## Supplementary material for "Development and Implementation of a scalable and versatile test for COVID-19 diagnostics in rural communities": Sup_Table_VII

Group 1 - Jan 2020 - Jun 2020

| A | B | C | D | E | F | G | H | I | J | K | L |
|---|---|---|---|---|---|---|---|---|---|---|---|
| Database | Database ID | GISAID ID | SK_N forward GCTGCAATCGTGCTACAACT | SK_N reverse TGAACTGTTGCGACTACGTG | SK_E forward TTCGGAAGAGACAGGTACGTT | SK_E reverse CACACAATCGAAGCGCAGTA | SK_S forward GCTGGTGCTGCAGCTTATTA | SK_S reverse AGGGTCAAGTGCACAGTCTA | 3' mismatches? | Multiple mismatches? | More than 1 primer set? |
| A | 1 | hCoV-19/Australia/NSW14/2020\|EPI_ISL_413600\|2020-0 | ···················· | ···················· | ···················· | ···················· | ···················· | ···················· | | | |
| A | 2 | hCoV-19/USA/WA13-UW9/2020\|EPI_ISL_413601\|2020-03-0 | ···················· | ···················· | ···················· | ···················· | ···················· | ···················· | | | |
| A | 3 | hCoV-19/USA/WA-S47/2020\|EPI_ISL_417100\|2020-02-29 | ···················· | ···················· | ···················· | ···················· | ···················· | ···················· | | | |
| A | 4 | hCoV-19/USA/WA-S48/2020\|EPI_ISL_417101\|2020-02-29 | ···················· | ···················· | ···················· | ···················· | ···················· | ···················· | | | |
| A | 5 | hCoV-19/USA/WA-S49/2020\|EPI_ISL_417102\|2020-03-04 | ···················· | ···················· | ···················· | ···················· | ···················· | ···················· | | | |
| A | 6 | hCoV-19/USA/WA-S77/2020\|EPI_ISL_417130\|2020-03-05 | ···················· | ···················· | ···················· | ···················· | ···················· | ···················· | | | |
| A | 7 | hCoV-19/USA/WA-S79/2020\|EPI_ISL_417132\|2020-03-05 | ···················· | ···················· | ···················· | ···················· | ···················· | ···················· | | | |
| A | 8 | hCoV-19/USA/WA-S80/2020\|EPI_ISL_417133\|2020-03-05 | ···················· | ···················· | ···················· | ···················· | ···················· | ···················· | | | |
| A | 9 | hCoV-19/USA/WA-S81/2020\|EPI_ISL_417134\|2020-02-26 | ···················· | ···················· | ···················· | ···················· | ···················· | ···················· | | | |
| A | 10 | hCoV-19/USA/WA-S82/2020\|EPI_ISL_417135\|2020-02-22 | ···················· | ···················· | ···················· | ···················· | ···················· | ···················· | | | |
| A | 11 | hCoV-19/USA/WA-S83/2020\|EPI_ISL_417136\|2020-03-05 | ···················· | ···················· | ···················· | ···················· | ···················· | ···················· | | | |
| A | 12 | hCoV-19/USA/WA-S84/2020\|EPI_ISL_417137\|2020-02-21 | ···················· | ···················· | ···················· | ···················· | ···················· | ···················· | | | |
| A | 13 | hCoV-19/USA/WA-S86/2020\|EPI_ISL_417139\|2020-03-01 | ···················· | ···················· | ···················· | ···················· | ···················· | ···················· | | | |
| A | 14 | hCoV-19/USA/WA-S87/2020\|EPI_ISL_417140\|2020-03-01 | ···················· | ···················· | ···················· | ···················· | ···················· | ···················· | | | |
| A | 15 | hCoV-19/USA/WA-S88/2020\|EPI_ISL_417141\|2020-03-01 | ···················· | ···················· | ···················· | ···················· | ···················· | ···················· | | | |
| A | 16 | hCoV-19/USA/WA-S89/2020\|EPI_ISL_417142\|2020-02-29 | ···················· | ···················· | ···················· | ···················· | ···················· | ···················· | | | |
| A | 17 | hCoV-19/USA/WA-S90/2020\|EPI_ISL_417143\|2020-02-29 | ···················· | ···················· | ···················· | ···················· | ···················· | ···················· | | | |
| A | 18 | hCoV-19/USA/WA-S91/2020\|EPI_ISL_417144\|2020-03-02 | ···················· | ···················· | ···················· | ···················· | ···················· | ···················· | | | |
| A | 19 | hCoV-19/USA/WA-S92/2020\|EPI_ISL_417145\|2020-02-29 | ···················· | ···················· | ···················· | ···················· | ···················· | ···················· | | | |
| A | 20 | hCoV-19/USA/WA-S93/2020\|EPI_ISL_417146\|2020-02-29 | ···················· | ···················· | ···················· | ···················· | ···················· | ···················· | | | |
| A | 21 | hCoV-19/USA/WA-S94/2020\|EPI_ISL_417147\|2020-02-28 | ···················· | ···················· | ···················· | ···················· | ···················· | ···················· | | | |
| A | 22 | hCoV-19/USA/WA-S95/2020\|EPI_ISL_417148\|2020-02-28 | ···················· | ···················· | ···················· | ···················· | ···················· | ···················· | | | |
| A | 23 | hCoV-19/USA/WA-S96/2020\|EPI_ISL_417149\|2020-02-28 | ···················· | ···················· | ···················· | ···················· | ···················· | ···················· | | | |
| A | 24 | hCoV-19/USA/WA-S97/2020\|EPI_ISL_417150\|2020-02-28 | ···················· | ···················· | ···················· | ···················· | ···················· | ···················· | | | |
| A | 25 | hCoV-19/USA/WA-S98/2020\|EPI_ISL_417151\|2020-02-29 | ···················· | ···················· | ···················· | ···················· | ···················· | ···················· | | | |
| A | 26 | hCoV-19/USA/WA-S99/2020\|EPI_ISL_417152\|2020-02-28 | ···················· | ···················· | ···················· | ···················· | ···················· | ···················· | | | |
| A | 27 | hCoV-19/USA/WA-S100/2020\|EPI_ISL_417153\|2020-02-29 | ···················· | ···················· | ···················· | ···················· | ···················· | ···················· | | | |
| A | 28 | hCoV-19/USA/WA-S101/2020\|EPI_ISL_417154\|2020-02-28 | ···················· | ···················· | ···················· | ···················· | ···················· | ···················· | | | |
| A | 29 | hCoV-19/USA/WA-S102/2020\|EPI_ISL_417155\|2020-02-28 | ···················· | ···················· | ···················· | ···················· | ···················· | ···················· | | | |
| A | 30 | hCoV-19/USA/WA-S103/2020\|EPI_ISL_417156\|2020-02-28 | ···················· | ···················· | ···················· | ···················· | ···················· | ···················· | | | |
| A | 31 | hCoV-19/USA/WA-S104/2020\|EPI_ISL_417157\|2020-03-05 | ···················· | ···················· | ···················· | ···················· | ···················· | ···················· | | | |
| A | 32 | hCoV-19/USA/WA-S105/2020\|EPI_ISL_417158\|2020-02-28 | ···················· | ···················· | ···················· | ···················· | ···················· | ···················· | | | |
| A | 33 | hCoV-19/USA/WA-S106/2020\|EPI_ISL_417159\|2020-02-29 | ···················· | ···················· | ···················· | ···················· | ···················· | ···················· | | | |
| A | 34 | hCoV-19/USA/WA-S107/2020\|EPI_ISL_417160\|2020-02-29 | ···················· | ···················· | ···················· | ···················· | ···················· | ···················· | | | |
| A | 35 | hCoV-19/USA/WA-S108/2020\|EPI_ISL_417161\|2020-02-29 | ···················· | ···················· | ···················· | ···················· | ···················· | ···················· | | | |
| A | 36 | hCoV-19/USA/WA-S109/2020\|EPI_ISL_417162\|2020-03-01 | ···················· | ···················· | ···················· | ···················· | ···················· | ···················· | | | |
| A | 37 | hCoV-19/USA/WA-S110/2020\|EPI_ISL_417163\|2020-03-05 | ···················· | ···················· | ···················· | ···················· | ···················· | ···················· | | | |
| A | 38 | hCoV-19/USA/WA-S111/2020\|EPI_ISL_417164\|2020-03-07 | ···················· | ···················· | ···················· | ···················· | ···················· | ···················· | | | |
| A | 39 | hCoV-19/USA/WA-S112/2020\|EPI_ISL_417165\|2020-03-08 | ···················· | ···················· | ···················· | ···················· | ···················· | ···················· | | | |
| A | 40 | hCoV-19/USA/WA-S113/2020\|EPI_ISL_417166\|2020-02-29 | ···················· | ···················· | ···················· | ···················· | ···················· | ···················· | | | |
| A | 41 | hCoV-19/USA/WA-S114/2020\|EPI_ISL_417167\|2020-03-05 | ···················· | ···················· | ···················· | ···················· | ···················· | ···················· | | | |
| A | 42 | hCoV-19/USA/WA-S115/2020\|EPI_ISL_417168\|2020-02-29 | ···················· | ···················· | ···················· | ···················· | ···················· | ···················· | | | |
| A | 43 | hCoV-19/USA/WA-S116/2020\|EPI_ISL_417169\|2020-03-02 | ···················· | ···················· | ···················· | ···················· | ···················· | ···················· | | | |
| A | 44 | hCoV-19/USA/WA-S117/2020\|EPI_ISL_417170\|2020-03-02 | ···················· | ···················· | ···················· | ···················· | ···················· | ···················· | | | |
| A | 45 | hCoV-19/USA/WA-S118/2020\|EPI_ISL_417171\|2020-03-01 | ···················· | ···················· | ···················· | ···················· | ···················· | ···················· | | | |
| A | 46 | hCoV-19/USA/WA-S119/2020\|EPI_ISL_417172\|2020-02-29 | ···················· | ···················· | ···················· | ···················· | ···················· | ···················· | | | |
| A | 47 | hCoV-19/USA/WA-S122/2020\|EPI_ISL_417175\|2020-03-02 | ···················· | ···················· | ···················· | ···················· | ···················· | ···················· | | | |
| A | 48 | hCoV-19/Hong Kong/HKPU1_2101/2020\|EPI_ISL_417176\|2 | ···················· | ···················· | ···················· | ···················· | ···················· | ···················· | | | |
| A | 49 | hCoV-19/Hong Kong/HKPU6_2101/2020\|EPI_ISL_417178\|2 | ···················· | ···················· | ···················· | ···················· | ···················· | ···················· | | | |
| A | 50 | hCoV-19/Hong Kong/HKPU19_0402/2020\|EPI_ISL_417181\| | ···················· | ···················· | ···················· | ···················· | ···················· | ···················· | | | |
| A | 51 | hCoV-19/Hong Kong/HKPU28_3001/2020\|EPI_ISL_417185\| | ···················· | ···················· | ···················· | ···················· | ···················· | ···················· | | | |
| A | 52 | hCoV-19/South Africa/R03006/2020\|EPI_ISL_417186\|20 | ···················· | ···················· | ···················· | ···················· | ···················· | ···················· | | | |
| A | 53 | hCoV-19/Hong Kong/HKPU29_0102/2020\|EPI_ISL_417187\| | ···················· | ···················· | ···················· | ···················· | ···················· | ···················· | | | |
| A | 54 | hCoV-19/Hong Kong/HKPU30_2901/2020\|EPI_ISL_417188\| | ···················· | ···················· | ···················· | ···················· | ···················· | ···················· | | | |
| A | 55 | hCoV-19/USA/MN5-MDH5/2020\|EPI_ISL_417191\|2020-03-1 | ···················· | ···················· | ···················· | ···················· | ···················· | ···················· | | | |
| A | 56 | hCoV-19/USA/MN25-MDH25/2020\|EPI_ISL_417192\|2020-03 | ···················· | ···················· | ···················· | ···················· | ···················· | N··················· | | | |
| A | 57 | hCoV-19/Hong Kong/HKPU32_0402/2020\|EPI_ISL_417193\| | ···················· | ···················· | ···················· | ···················· | ···················· | ···················· | | | |
| A | 58 | hCoV-19/USA/MN26-MDH26/2020\|EPI_ISL_417194\|2020-03 | ···················· | ···················· | ···················· | ···················· | ···················· | ···················· | | | |
| A | 59 | hCoV-19/USA/MN29-MDH29/2020\|EPI_ISL_417196\|2020-03 | ···················· | ···················· | ···················· | ···················· | ···················· | ···················· | | | |
| A | 60 | hCoV-19/Hong Kong/HKPU34_3001/2020\|EPI_ISL_417197\| | ···················· | ···················· | ···················· | ···················· | ···················· | ···················· | | | |
| A | 61 | hCoV-19/Finland/13M83/2020\|EPI_ISL_418400\|2020-03- | ···················· | ···················· | ···················· | ···················· | ···················· | ···················· | | | |
| A | 62 | hCoV-19/Finland/14M12/2020\|EPI_ISL_418401\|2020-03- | ···················· | ···················· | ···················· | ···················· | ···················· | ···················· | | | |
| A | 63 | hCoV-19/Finland/14M13/2020\|EPI_ISL_418402\|2020-03- | ···················· | ···················· | ···················· | ···················· | ···················· | ···················· | | | |
| A | 64 | hCoV-19/Finland/14M14/2020\|EPI_ISL_418403\|2020-03- | ···················· | ···················· | ···················· | ···················· | ···················· | ···················· | | | |
| A | 65 | hCoV-19/Finland/14M16/2020\|EPI_ISL_418404\|2020-03- | ···················· | ···················· | ···················· | ···················· | ···················· | ···················· | | | |
| A | 66 | hCoV-19/Finland/14M20/2020\|EPI_ISL_418405\|2020-03- | ···················· | ···················· | ···················· | ···················· | ···················· | ···················· | | | |
| A | 67 | hCoV-19/Finland/14M26/2020\|EPI_ISL_418406\|2020-03- | ···················· | ···················· | ···················· | ···················· | ···················· | ···················· | | | |
| A | 68 | hCoV-19/Finland/14M32/2020\|EPI_ISL_418408\|2020-03- | ···················· | ···················· | ···················· | ···················· | ···················· | ···················· | | | |

```
A    69 hCoV-19/Finland/14M74/2020|EPI_ISL_418409|2020-03-    ···················· ···················· ···················· ···················· ···················· ····················
A    70 hCoV-19/Finland/14M77/2020|EPI_ISL_418410|2020-03-    ···················· ···················· ···················· ···················· ···················· ····················
A    71 hCoV-19/Finland/14M82/2020|EPI_ISL_418411|2020-03-    ···················· ···················· ···················· ···················· ···················· ····················
A    72 hCoV-19/France/ARA-094100/2020|EPI_ISL_418412|2020    ···················· ···················· ···················· ···················· ···················· ····················
A    73 hCoV-19/France/ARA-09428/2020|EPI_ISL_418413|2020-    ···················· ···················· ···················· ···················· ···················· ····················
A    74 hCoV-19/France/ARA-09434/2020|EPI_ISL_418414|2020-    ···················· ···················· ···················· ···················· ···················· ····················
A    75 hCoV-19/France/ARA-09588/2020|EPI_ISL_418416|2020-    ···················· ···················· ···················· ···················· ···················· ····················
A    76 hCoV-19/France/ARA-09686/2020|EPI_ISL_418417|2020-    ···················· ···················· ···················· ···················· ···················· ····················
A    77 hCoV-19/France/ARA-10163/2020|EPI_ISL_418418|2020-    ···················· ···················· ···················· ···················· ···················· ····················
A    78 hCoV-19/France/ARA-10165/2020|EPI_ISL_418419|2020-    ···················· ···················· ···················· ···················· ···················· ····················
A    79 hCoV-19/France/ARA-10910/2020|EPI_ISL_418430|2020-    ···················· ···················· ···················· ···················· ···················· ····················
A    81 hCoV-19/France/ARA-11036/2020|EPI_ISL_418432|2020-    ···················· ···················· ···················· ···················· ···················· ····················
A    83 hCoV-19/Spain/CT-ISCIII-201397/2020|EPI_ISL_419707    ···················· ···················· ···················· ···················· ···················· ····················
A    84 hCoV-19/Spain/PV-ISCIII-201602/2020|EPI_ISL_419709    ···················· ···················· ···················· ···················· ···················· ····················
A    85 hCoV-19/USA/VA-DCLS-0016/2020|EPI_ISL_419710|2020-    ···················· ···················· ···················· ···················· ···················· ··A·················
A    86 hCoV-19/USA/VA-DCLS-0017/2020|EPI_ISL_419711|2020-    ···················· ···················· ···················· ···················· ···················· ····················
A    87 hCoV-19/USA/VA-DCLS-0020/2020|EPI_ISL_419712|2020-    ···················· ···················· ···················· ···················· ···················· ····················
A    88 hCoV-19/USA/VA-DCLS-0021/2020|EPI_ISL_419713|2020-    ···················· ···················· ···················· ···················· ···················· ····················
A    89 hCoV-19/Australia/VIC108/2020|EPI_ISL_419714|2020-    ···················· ···················· ···················· ···················· ···················· ····················
A    90 hCoV-19/Australia/VIC111/2020|EPI_ISL_419715|2020-    ···················· ···················· ···················· ···················· ···················· ····················
A    91 hCoV-19/Australia/VIC110/2020|EPI_ISL_419716|2020-    ···················· ···················· ···················· ···················· ···················· ····················
A    92 hCoV-19/Australia/VIC115/2020|EPI_ISL_419717|2020-    ···················· ···················· ···················· ···················· ···················· ····················
A    93 hCoV-19/Australia/VIC109/2020|EPI_ISL_419718|2020-    ···················· ···················· ···················· ···················· ···················· ····················
A    94 hCoV-19/Australia/VIC118/2020|EPI_ISL_419719|2020-    ···················· ···················· ···················· ···················· ···················· ····················
A    95 hCoV-19/Australia/VIC107/2020|EPI_ISL_419720|2020-    ···················· ···················· ···················· ···················· ···················· ····················
A    96 hCoV-19/Australia/VIC116/2020|EPI_ISL_419721|2020-    ···················· ···················· ···················· ···················· ···················· ····················
A    97 hCoV-19/Australia/VIC120/2020|EPI_ISL_419722|2020-    ···················· ···················· ···················· ···················· ···················· ····················
A    98 hCoV-19/Australia/VIC121/2020|EPI_ISL_419723|2020-    ···················· ···················· ···················· ···················· ···················· ····················
A    99 hCoV-19/Australia/VIC114/2020|EPI_ISL_419725|2020-    ···················· ···················· ···················· ···················· ···················· ····················
A   100 hCoV-19/Australia/VIC119/2020|EPI_ISL_419726|2020-    ···················· ···················· ···················· ···················· ···················· ····················
A   101 hCoV-19/Australia/VIC117/2020|EPI_ISL_419727|2020-    ···················· ···················· ···················· ···················· ···················· ····················
A   102 hCoV-19/Australia/VIC126/2020|EPI_ISL_419728|2020-    ···················· ···················· ···················· ···················· ···················· ····················
A   103 hCoV-19/Australia/VIC127/2020|EPI_ISL_419729|2020-    ···················· ···················· ···················· ···················· ···················· ····················
A   104 hCoV-19/Australia/VIC128/2020|EPI_ISL_419730|2020-    ···················· ···················· ···················· ···················· ···················· ····················
A   105 hCoV-19/Australia/VIC123/2020|EPI_ISL_419731|2020-    ···················· ···················· ···················· ···················· ···················· ····················
A   106 hCoV-19/Australia/VIC129/2020|EPI_ISL_419732|2020-    ···················· ···················· ···················· ···················· ···················· ····················
A   107 hCoV-19/Australia/VIC13/2020|EPI_ISL_419733|2020-0    ···················· ···················· ···················· ···················· ···················· ····················
A   108 hCoV-19/Australia/VIC17/2020|EPI_ISL_419736|2020-0    ···················· ···················· ···················· ···················· ···················· ····················
A   109 hCoV-19/Australia/VIC18/2020|EPI_ISL_419737|2020-0    ···················· ···················· ···················· ···················· ···················· ····················
A   110 hCoV-19/Australia/VIC24/2020|EPI_ISL_419739|2020-0    ···················· ···················· ···················· ···················· ···················· ····················
A   111 hCoV-19/Australia/VIC21/2020|EPI_ISL_419740|2020-0    ···················· ···················· ···················· ···················· ···················· ····················
A   112 hCoV-19/Australia/VIC20/2020|EPI_ISL_419741|2020-0    ···················· ···················· ···················· ···················· ···················· ····················
A   113 hCoV-19/Australia/VIC22/2020|EPI_ISL_419742|2020-0    ···················· ···················· ···················· ···················· ···················· ····················
A   114 hCoV-19/Australia/VIC23/2020|EPI_ISL_419743|2020-0    ···················· ···················· ···················· ···················· ···················· ····················
A   115 hCoV-19/Australia/VIC25/2020|EPI_ISL_419744|2020-0    ···················· ···················· ···················· ···················· ···················· ····················
A   116 hCoV-19/Australia/VIC26/2020|EPI_ISL_419745|2020-0    ···················· ···················· ···················· ···················· ···················· ····················
A   117 hCoV-19/Australia/VIC27/2020|EPI_ISL_419746|2020-0    ···················· ···················· ···················· ···················· ···················· ····················
A   118 hCoV-19/Australia/VIC28/2020|EPI_ISL_419747|2020-0    ···················· ···················· ···················· ···················· ···················· ····················
A   119 hCoV-19/Australia/VIC29/2020|EPI_ISL_419748|2020-0    ···················· ···················· ···················· ···················· ···················· ····················
A   120 hCoV-19/Australia/VIC30/2020|EPI_ISL_419749|2020-0    ···················· ···················· ···················· ···················· ···················· ····················
A   121 hCoV-19/Australia/VIC41/2020|EPI_ISL_419760|2020-0    ···················· ···················· ···················· ···················· ···················· ····················
A   122 hCoV-19/Australia/VIC42/2020|EPI_ISL_419761|2020-0    ···················· ···················· ···················· ···················· ···················· ····················
A   123 hCoV-19/Australia/VIC48/2020|EPI_ISL_419762|2020-0    ···················· ···················· ···················· ···················· ···················· ····················
A   124 hCoV-19/Australia/VIC43/2020|EPI_ISL_419763|2020-0    ···················· ···················· ···················· ···················· ···················· ····················
A   125 hCoV-19/Australia/VIC49/2020|EPI_ISL_419764|2020-0    ···················· ···················· ···················· ···················· ···················· ····················
A   126 hCoV-19/Australia/VIC52/2020|EPI_ISL_419768|2020-0    ···················· ···················· ···················· ···················· ···················· ····················
A   127 hCoV-19/Australia/VIC78/2020|EPI_ISL_419790|2020-0    ···················· ···················· ···················· ···················· ···················· ····················
A   128 hCoV-19/Australia/VIC79/2020|EPI_ISL_419791|2020-0    ···················· ···················· ···················· ···················· ···················· ····················
A   129 hCoV-19/Australia/VIC80/2020|EPI_ISL_419792|2020-0    ···················· ···················· ···················· ···················· ···················· ····················
A   130 hCoV-19/Australia/VIC81/2020|EPI_ISL_419793|2020-0    ···················· ···················· ···················· ···················· ···················· ····················
A   131 hCoV-19/Australia/VIC82/2020|EPI_ISL_419794|2020-0    ···················· ···················· ···················· ···················· ···················· ····················
A   132 hCoV-19/Australia/VIC85/2020|EPI_ISL_419795|2020-0    ···················· ···················· ···················· ···················· ···················· ····················
A   133 hCoV-19/Australia/VIC86/2020|EPI_ISL_419796|2020-0    ···················· ···················· ···················· ···················· ···················· ····················
A   134 hCoV-19/Australia/VIC87/2020|EPI_ISL_419797|2020-0    ···················· ···················· ···················· ···················· ···················· ····················
A   135 hCoV-19/Australia/VIC88/2020|EPI_ISL_419798|2020-0    ···················· ···················· ···················· ···················· ···················· ····················
A   137 hCoV-19/England/20139053304/2020|EPI_ISL_420707|20    ···················· ···················· ···················· ···················· ···················· ····················
A   138 hCoV-19/England/20139053504/2020|EPI_ISL_420708|20    ···················· ···················· ···················· ···················· ···················· ····················
A   140 hCoV-19/England/20139055304/2020|EPI_ISL_420715|20    ···················· ···················· ···················· ···················· ···················· ····················
A   141 hCoV-19/England/20139056904/2020|EPI_ISL_420716|20    ···················· ···················· ···················· ···················· ···················· ····················
A   143 hCoV-19/England/20139057204/2020|EPI_ISL_420718|20    ···················· ···················· ···················· ···················· ···················· ····················
A   144 hCoV-19/England/20139057404/2020|EPI_ISL_420719|20    ···················· ···················· ···················· ···················· ···················· ····················
A   146 hCoV-19/England/20139057604/2020|EPI_ISL_420721|20    ···················· ···················· ···················· ···················· ···················· ····················
A   147 hCoV-19/England/20139057704/2020|EPI_ISL_420722|20    ···················· ···················· ···················· ···················· ···················· ····················
A   148 hCoV-19/England/20139058104/2020|EPI_ISL_420723|20    ···················· ···················· ···················· ···················· ···················· ····················
A   149 hCoV-19/England/20139058304/2020|EPI_ISL_420724|20    ···················· ···················· ···················· ···················· ···················· ····················
A   150 hCoV-19/England/20139059004/2020|EPI_ISL_420727|20    ···················· ···················· ···················· ···················· ···················· ····················
A   151 hCoV-19/England/20139059504/2020|EPI_ISL_420728|20    ···················· ···················· ···················· ···················· ···················· ····················
A   153 hCoV-19/England/20140038604/2020|EPI_ISL_420750|20    ···················· ···················· ···················· ···················· ···················· ····················
```

| | | | |
|---|---|---|---|
| A | 154 | hCoV-19/England/20140038704/2020\|EPI_ISL_420751\|20 | .................... .................... .................... .................... .................... .................... |
| A | 155 | hCoV-19/England/20142062604/2020\|EPI_ISL_420774\|20 | .................... .................... .................... .................... .................... .................... |
| A | 157 | hCoV-19/USA/AZ_4811/2020\|EPI_ISL_420784\|2020-03-02 | .................... .................... .................... .................... .................... .................... |
| A | 158 | hCoV-19/USA/FL_6318/2020\|EPI_ISL_420785\|2020-03-02 | .................... .................... .................... .................... .................... .................... |
| A | 159 | hCoV-19/USA/GA_1320/2020\|EPI_ISL_420786\|2020-03-03 | .................... .................... .................... .................... .................... .................... |
| A | 160 | hCoV-19/USA/GA_1299/2020\|EPI_ISL_420787\|2020-03-03 | .................... .................... .................... .................... .................... .................... |
| A | 161 | hCoV-19/USA/GA_1445/2020\|EPI_ISL_420788\|2020-03-04 | .................... .................... .................... .................... .................... .................... |
| A | 162 | hCoV-19/USA/IL_1375/2020\|EPI_ISL_420789\|2020-03-01 | .................... .................... .................... .................... .................... .................... |
| A | 163 | hCoV-19/USA/IL_1293/2020\|EPI_ISL_420790\|2020-03-01 | .................... .................... .................... .................... .................... .................... |
| A | 164 | hCoV-19/USA/NH_0004/2020\|EPI_ISL_420791\|2020-02-29 | .................... .................... .................... .................... .................... .................... |
| A | 165 | hCoV-19/USA/NH_0008/2020\|EPI_ISL_420792\|2020-03-02 | .................... .................... .................... .................... .................... .................... |
| A | 166 | hCoV-19/USA/NY_2929/2020\|EPI_ISL_420793\|2020-03-02 | .................... .................... .................... .................... .................... .................... |
| A | 167 | hCoV-19/USA/OR_5430/2020\|EPI_ISL_420794\|2020-03-01 | .................... .................... .................... .................... .................... .................... |
| A | 168 | hCoV-19/USA/RI_0556/2020\|EPI_ISL_420795\|2020-03-01 | .................... .................... .................... .................... .................... .................... |
| A | 169 | hCoV-19/USA/TX_2039/2020\|EPI_ISL_420796\|2020-02-29 | .................... .................... .................... .................... .................... .................... |
| A | 170 | hCoV-19/USA/TX_2817/2020\|EPI_ISL_420797\|2020-03-01 | .................... .................... .................... .................... .................... .................... |
| A | 171 | hCoV-19/USA/TX_2967/2020\|EPI_ISL_420798\|2020-03-01 | .................... .................... .................... .................... .................... .................... |
| A | 172 | hCoV-19/South Korea/BA-ACH_2604/2020\|EPI_ISL_42079 | .................... .................... .................... .................... .................... .................... |
| A | 175 | hCoV-19/Australia/NT17/2020\|EPI_ISL_426904\|2020 | .................... .................... .................... .................... .................... .................... |
| A | 179 | hCoV-19/Sweden/20-08142/2020\|EPI_ISL_429115\|2020-0 | .................... .................... .................... .................... .................... .................... |
| A | 180 | hCoV-19/Sweden/20-08143/2020\|EPI_ISL_429116\|2020-0 | .................... .................... .................... .................... .................... .................... |
| A | 181 | hCoV-19/Sweden/20-08145/2020\|EPI_ISL_429117\|2020-0 | .................... .................... .................... .................... .................... .................... |
| A | 182 | hCoV-19/Sweden/20-08146/2020\|EPI_ISL_429118\|2020-0 | .................... .................... .................... .................... .................... .................... |
| A | 183 | hCoV-19/Sweden/20-08148/2020\|EPI_ISL_429119\|2020-0 | .................... .................... .................... .................... .................... .................... |
| A | 184 | hCoV-19/Sweden/20-08149/2020\|EPI_ISL_429120\|2020-0 | .................... .................... .................... .................... .................... .................... |
| A | 185 | hCoV-19/Sweden/20-08151/2020\|EPI_ISL_429121\|2020-0 | .................... .................... .................... .................... .................... .................... |
| A | 186 | hCoV-19/Sweden/20-08155/2020\|EPI_ISL_429122\|2020-0 | .................... .................... .................... .................... .................... .................... |
| A | 187 | hCoV-19/Sweden/20-08169/2020\|EPI_ISL_429123\|2020-0 | .................... .................... .................... .................... .................... .................... |
| A | 188 | hCoV-19/Sweden/20-08170/2020\|EPI_ISL_429124\|2020-0 | .................... .................... .................... .................... .................... .................... |
| A | 189 | hCoV-19/Sweden/20-08173/2020\|EPI_ISL_429125\|2020-0 | .................... .................... .................... .................... .................... .................... |
| A | 190 | hCoV-19/Sweden/20-08320/2020\|EPI_ISL_429126\|2020-0 | .................... .................... .................... .................... .................... .................... |
| A | 191 | hCoV-19/Sweden/20-08332/2020\|EPI_ISL_429127\|2020-0 | .................... .................... .................... .................... .................... .................... |
| A | 192 | hCoV-19/Sweden/20-08680/2020\|EPI_ISL_429128\|2020-0 | .................... .................... .................... .................... .................... .................... |
| A | 193 | hCoV-19/Sweden/20-08681/2020\|EPI_ISL_429129\|2020-0 | .................... .................... .................... .................... .................... .................... |
| A | 194 | hCoV-19/Sweden/20-08683/2020\|EPI_ISL_429130\|2020-0 | .................... .................... .................... .................... .................... .................... |
| A | 195 | hCoV-19/Sweden/20-08684/2020\|EPI_ISL_429131\|2020-0 | .................... .................... .................... .................... .................... .................... |
| A | 196 | hCoV-19/Sweden/20-08685/2020\|EPI_ISL_429132\|2020-0 | .................... .................... .................... .................... .................... .................... |
| A | 197 | hCoV-19/Sweden/20-08690/2020\|EPI_ISL_429133\|2020-0 | .................... .................... .................... .................... .................... .................... |
| A | 198 | hCoV-19/Sweden/20-08693/2020\|EPI_ISL_429134\|2020-0 | .................... .................... .................... .................... .................... .................... |
| A | 199 | hCoV-19/Sweden/20-08719/2020\|EPI_ISL_429135\|2020-0 | .................... .................... .................... .................... .................... .................... |
| A | 200 | hCoV-19/Sweden/20-50048/2020\|EPI_ISL_429136\|2020-0 | .................... .................... .................... .................... .................... .................... |
| A | 201 | hCoV-19/Sweden/20-50076/2020\|EPI_ISL_429137\|2020-0 | .................... .................... .................... .................... .................... .................... |
| A | 202 | hCoV-19/Sweden/20-50087/2020\|EPI_ISL_429138\|2020-0 | .................... .................... .................... .................... .................... .................... |
| A | 203 | hCoV-19/Sweden/20-50123/2020\|EPI_ISL_429139\|2020-0 | .................... .................... .................... .................... .................... .................... |
| A | 204 | hCoV-19/Sweden/20-50125/2020\|EPI_ISL_429140\|2020-0 | .................... .................... .................... .................... .................... .................... |
| A | 205 | hCoV-19/Sweden/20-50130/2020\|EPI_ISL_429141\|2020-0 | .................... .................... .................... .................... .................... .................... |
| A | 206 | hCoV-19/Sweden/20-50131/2020\|EPI_ISL_429142\|2020-0 | .................... .................... .................... .................... .................... .................... |
| A | 207 | hCoV-19/Sweden/20-50132/2020\|EPI_ISL_429143\|2020-0 | .................... .................... .................... .................... .................... .................... |
| A | 208 | hCoV-19/Sweden/20-50133/2020\|EPI_ISL_429144\|2020-0 | .................... .................... .................... .................... .................... .................... |
| A | 209 | hCoV-19/Sweden/20-50170/2020\|EPI_ISL_429145\|2020-0 | .................... .................... .................... .................... .................... .................... |
| A | 210 | hCoV-19/Sweden/20-50171/2020\|EPI_ISL_429146\|2020-0 | .................... .................... .................... .................... .................... .................... |
| A | 211 | hCoV-19/Sweden/20-50172/2020\|EPI_ISL_429147\|2020-0 | .................... .................... .................... .................... .................... .................... |
| A | 212 | hCoV-19/Sweden/20-50173/2020\|EPI_ISL_429148\|2020-0 | .................... .................... .................... .................... .................... .................... |
| A | 213 | hCoV-19/Sweden/20-50175/2020\|EPI_ISL_429150\|2020-0 | .................... .................... .................... .................... .................... .................... |
| A | 214 | hCoV-19/Sweden/20-50176/2020\|EPI_ISL_429151\|2020-0 | .................... .................... .................... .................... .................... .................... |
| A | 215 | hCoV-19/Sweden/20-50179/2020\|EPI_ISL_429152\|2020-0 | .................... .................... .................... .................... .................... .................... |
| A | 216 | hCoV-19/Sweden/20-50180/2020\|EPI_ISL_429153\|2020-0 | .................... .................... .................... .................... .................... .................... |
| A | 217 | hCoV-19/Sweden/20-50181/2020\|EPI_ISL_429154\|2020-0 | .................... .................... .................... .................... .................... .................... |
| A | 218 | hCoV-19/Sweden/20-50230/2020\|EPI_ISL_429155\|2020-0 | .................... .................... .................... .................... .................... .................... |
| A | 219 | hCoV-19/Sweden/20-50232/2020\|EPI_ISL_429156\|2020-0 | .................... .................... .................... .................... .................... .................... |
| A | 220 | hCoV-19/Sweden/20-50234/2020\|EPI_ISL_429157\|2020-0 | .................... .................... .................... .................... .................... .................... |
| A | 221 | hCoV-19/Sweden/20-50235/2020\|EPI_ISL_429158\|2020-0 | .................... .................... .................... .................... .................... .................... |
| A | 222 | hCoV-19/Sweden/20-50312/2020\|EPI_ISL_429160\|2020-0 | .................... .................... .................... .................... .................... .................... |
| A | 223 | hCoV-19/Sweden/20-50315/2020\|EPI_ISL_429161\|2020-0 | .................... .................... .................... .................... .................... .................... |
| A | 224 | hCoV-19/Sweden/20-50357/2020\|EPI_ISL_429162\|2020-0 | .................... .................... .................... .................... .................... .................... |
| A | 225 | hCoV-19/Sweden/20-50361/2020\|EPI_ISL_429163\|2020-0 | .................... .................... .................... .................... .................... .................... |
| A | 226 | hCoV-19/Thailand/Bangkok-0025/2020\|EPI_ISL_429164\| | .................... .................... .................... .................... .................... .................... |
| A | 227 | hCoV-19/Thailand/Bangkok-0026/2020\|EPI_ISL_429165\| | .................... .................... .................... .................... .................... .................... |
| A | 228 | hCoV-19/Thailand/Bangkok-0028/2020\|EPI_ISL_429166\| | .................... .................... .................... .................... .................... .................... |
| A | 229 | hCoV-19/Thailand/Bangkok-0029/2020\|EPI_ISL_429167\| | .................... .................... .................... .................... .................... .................... |
| A | 230 | hCoV-19/Thailand/Bangkok-0033/2020\|EPI_ISL_429169\| | .................... .................... .................... .................... .................... .................... |
| A | 231 | hCoV-19/Thailand/Bangkok-0034/2020\|EPI_ISL_429170\| | .................... .................... .................... .................... .................... .................... |
| A | 232 | hCoV-19/Thailand/Bangkok-0035/2020\|EPI_ISL_429171\| | .................... .................... .................... .................... .................... .................... |
| A | 233 | hCoV-19/Thailand/Bangkok-0036/2020\|EPI_ISL_429172\| | .................... .................... .................... .................... .................... .................... |
| A | 234 | hCoV-19/Thailand/Bangkok-0037/2020\|EPI_ISL_429173\| | .................... .................... .................... .................... .................... .................... |
| A | 235 | hCoV-19/Thailand/Bangkok-0038/2020\|EPI_ISL_429174\| | .................... .................... .................... .................... .................... .................... |
| A | 236 | hCoV-19/Thailand/Bangkok-0039/2020\|EPI_ISL_429175\| | .................... .................... .................... .................... .................... .................... |
| A | 237 | hCoV-19/Thailand/Bangkok-0041/2020\|EPI_ISL_429177\| | .................... .................... .................... .................... .................... .................... |

```
A 238 hCoV-19/Thailand/Bangkok-0042/2020|EPI_ISL_429178| .................... .................... .................... .................... .................... ....................
A 239 hCoV-19/Thailand/Bangkok-0043/2020|EPI_ISL_429179| .................... .................... .................... .................... .................... ....................
A 241 hCoV-19/Thailand/Bangkok-0046/2020|EPI_ISL_429182| .................... .................... .................... .................... .................... ....................
A 242 hCoV-19/Thailand/Bangkok-0047/2020|EPI_ISL_429183| .................... .................... .................... .................... .................... ....................
A 243 hCoV-19/Thailand/Bangkok-0048/2020|EPI_ISL_429184| .................... .................... .................... .................... .................... ....................
A 244 hCoV-19/Russia/StPetersburg-RII6027S/2020|EPI_ISL_ .................... .................... .................... .................... .................... ....................
A 245 hCoV-19/Russia/StPetersburg-RII6031S/2020|EPI_ISL_ .................... .................... .................... .................... .................... ....................
A 246 hCoV-19/Russia/StPetersburg-RII6039S/2020|EPI_ISL_ .................... .................... .................... .................... .................... ....................
A 247 hCoV-19/Russia/StPetersburg-RII6043S/2020|EPI_ISL_ .................... .................... .................... .................... .................... ....................
A 248 hCoV-19/Russia/StPetersburg-RII6053S/2020|EPI_ISL_ .................... .................... .................... .................... .................... ....................
A 249 hCoV-19/Russia/StPetersburg-RII6055S/2020|EPI_ISL_ .................... .................... .................... .................... .................... ....................
A 250 hCoV-19/Russia/StPetersburg-RII6056S/2020|EPI_ISL_ .................... .................... .................... .................... .................... ....................
A 251 hCoV-19/Russia/StPetersburg-RII6060S/2020|EPI_ISL_ .................... .................... .................... .................... .................... ....................
A 252 hCoV-19/Russia/StPetersburg-RII6061S/2020|EPI_ISL_ .................... .................... .................... .................... .................... ....................
A 253 hCoV-19/Russia/StPetersburg-RII6063S/2020|EPI_ISL_ .................... .................... .................... .................... .................... ....................
A 254 hCoV-19/Russia/StPetersburg-RII6065S/2020|EPI_ISL_ .................... .................... .................... .................... .................... ....................
A 255 hCoV-19/Russia/StPetersburg-RII6086S/2020|EPI_ISL_ .................... .................... .................... .................... .................... ....................
A 256 hCoV-19/Russia/Ulan-Ude-RII4562V/2020|EPI_ISL_4301 .................... .................... .................... .................... .................... ....................
A 257 hCoV-19/Netherlands/Eindhoven_1363782/2020|EPI_ISL .................... .................... .................... .................... .................... ....................
A 258 hCoV-19/Netherlands/Haarlem_1363688/2020|EPI_ISL_4 .................... .................... .................... .................... .................... ....................
A 259 hCoV-19/Netherlands/Hardinxveld_Giessendam_1364806 .................... .................... .................... .................... .................... ....................
A 260 hCoV-19/Netherlands/Helmond_1363548/2020|EPI_ISL_4 .................... .................... .................... .................... .................... ....................
A 261 hCoV-19/Netherlands/Houten_1363498/2020|EPI_ISL_41 .................... .................... .................... .................... .................... ....................
A 262 hCoV-19/Netherlands/Naarden_1364774/2020|EPI_ISL_4 .................... .................... .................... .................... .................... ....................
A 263 hCoV-19/Netherlands/Nootdorp_1364222/2020|EPI_ISL_ .................... .................... .................... .................... .................... ....................
A 264 hCoV-19/Netherlands/Oisterwijk_1364072/2020|EPI_IS .................... .................... .................... .................... .................... ....................
A 265 hCoV-19/Netherlands/Rotterdam_1363790/2020|EPI_ISL .................... .................... .................... .................... .................... ....................
A 266 hCoV-19/Netherlands/Rotterdam_1364040/2020|EPI_ISL .................... .................... .................... .................... .................... ....................
A 267 hCoV-19/Netherlands/Rotterdam_1364740/2020|EPI_ISL .................... .................... .................... .................... .................... ....................
A 268 hCoV-19/Netherlands/Tilburg_1364286/2020|EPI_ISL_4 .................... .................... .................... .................... .................... ....................
A 269 hCoV-19/Netherlands/Utrecht_1363564/2020|EPI_ISL_4 .................... .................... .................... .................... .................... ....................
A 270 hCoV-19/Netherlands/Utrecht_1363628/2020|EPI_ISL_4 .................... .................... .................... .................... .................... ....................
A 271 hCoV-19/Netherlands/Utrecht_1364066/2020|EPI_ISL_4 .................... .................... .................... .................... .................... ....................
A 272 hCoV-19/Netherlands/Zeewolde_1365080/2020|EPI_ISL_ .................... .................... .................... .................... .................... ....................
A 273 hCoV-19/Taiwan/NTU03/2020|EPI_ISL_413592|2020-03-0 .................... .................... .................... .................... .................... ....................
A 274 hCoV-19/Luxembourg/Lux1/2020|EPI_ISL_413593|2020-0 .................... .................... .................... .................... .................... ....................
A 275 hCoV-19/Australia/NSW08/2020|EPI_ISL_413594|2020-0 .................... .................... .................... .................... .................... ....................
A 276 hCoV-19/Australia/NSW09/2020|EPI_ISL_413595|2020-0 .................... .................... .................... .................... .................... ....................
A 277 hCoV-19/Australia/NSW10/2020|EPI_ISL_413596|2020-0 .................... .................... .................... .................... .................... ....................
A 278 hCoV-19/Australia/NSW11/2020|EPI_ISL_413597|2020-0 .................... .................... .................... .................... .................... ....................
A 279 hCoV-19/Australia/NSW12/2020|EPI_ISL_413598|2020-0 .................... .................... .................... .................... .................... ....................
A 280 hCoV-19/Australia/NSW13/2020|EPI_ISL_413599|2020-0 .................... .................... .................... .................... .................... ....................
A 281 hCoV-19/USA/WA-S50/2020|EPI_ISL_417103|2020-03-05 .................... .................... .................... .................... .................... ....................
A 282 hCoV-19/USA/WA-S51/2020|EPI_ISL_417104|2020-03-03 .................... .................... .................... .................... .................... ....................
A 283 hCoV-19/USA/WA-S52/2020|EPI_ISL_417105|2020-03-03 .................... .................... .................... .................... .................... ....................
A 284 hCoV-19/USA/WA-S53/2020|EPI_ISL_417106|2020-03-03 .................... .................... .................... .................... .................... ....................
A 285 hCoV-19/USA/WA-S54/2020|EPI_ISL_417107|2020-03-05 .................... .................... .................... .................... .................... ....................
A 286 hCoV-19/USA/WA-S55/2020|EPI_ISL_417108|2020-02-29 .................... .................... .................... .................... .................... ....................
A 287 hCoV-19/USA/WA-S57/2020|EPI_ISL_417110|2020-03-03 .................... .................... .................... .................... .................... ....................
A 288 hCoV-19/USA/WA-S58/2020|EPI_ISL_417111|2020-03-05 .................... .................... .................... .................... .................... ....................
A 289 hCoV-19/USA/WA-S59/2020|EPI_ISL_417112|2020-03-02 .................... .................... .................... .................... .................... ....................
A 290 hCoV-19/USA/WA-S61/2020|EPI_ISL_417114|2020-03-05 .................... .................... .................... .................... .................... ....................
A 291 hCoV-19/USA/WA-S62/2020|EPI_ISL_417115|2020-03-03 .................... .................... .................... .................... .................... ....................
A 292 hCoV-19/USA/WA-S63/2020|EPI_ISL_417116|2020-03-04 .................... .................... .................... .................... .................... ....................
A 293 hCoV-19/USA/WA-S64/2020|EPI_ISL_417117|2020-03-03 .................... .................... .................... .................... .................... ....................
A 294 hCoV-19/USA/WA-S65/2020|EPI_ISL_417118|2020-03-03 .................... .................... .................... .................... .................... ....................
A 295 hCoV-19/USA/WA-S66/2020|EPI_ISL_417119|2020-03-06 .................... .................... .................... .................... .................... ....................
A 296 hCoV-19/USA/WA-S67/2020|EPI_ISL_417120|2020-03-04 .................... .................... .................... .................... .................... ....................
A 297 hCoV-19/USA/WA-S68/2020|EPI_ISL_417121|2020-03-04 .................... .................... .................... .................... .................... ....................
A 298 hCoV-19/USA/WA-S69/2020|EPI_ISL_417122|2020-03-05 .................... .................... .................... .................... .................... ....................
A 299 hCoV-19/USA/WA-S70/2020|EPI_ISL_417123|2020-03-05 .................... .................... .................... .................... .................... ....................
A 300 hCoV-19/USA/WA-S71/2020|EPI_ISL_417124|2020-03-05 .................... .................... .................... .................... .................... ....................
A 301 hCoV-19/USA/WA-S72/2020|EPI_ISL_417125|2020-03-06 .................... .................... .................... .................... .................... ....................
A 302 hCoV-19/USA/WA-S73/2020|EPI_ISL_417126|2020-03-05 .................... .................... .................... .................... .................... ....................
A 303 hCoV-19/USA/WA-S74/2020|EPI_ISL_417127|2020-03-05 .................... .................... .................... .................... .................... ....................
A 304 hCoV-19/USA/WA-S75/2020|EPI_ISL_417128|2020-03-05 .................... .................... .................... .................... .................... ....................
A 305 hCoV-19/USA/WA-S76/2020|EPI_ISL_417129|2020-03-05 .................... .................... .................... .................... .................... ....................
A 306 hCoV-19/France/ARA-10170/2020|EPI_ISL_418420|2020- .................... .................... .................... .................... .................... ....................
A 307 hCoV-19/France/ARA-10172/2020|EPI_ISL_418421|2020- .................... .................... .................... .................... .................... ....................
A 308 hCoV-19/France/ARA-10184/2020|EPI_ISL_418422|2020- .................... .................... .................... .................... .................... ....................
A 309 hCoV-19/France/ARA-10188/2020|EPI_ISL_418423|2020- .................... .................... .................... .................... .................... ....................
A 310 hCoV-19/France/ARA-10189/2020|EPI_ISL_418424|2020- .................... .................... .................... .................... .................... ....................
A 311 hCoV-19/France/ARA-10192/2020|EPI_ISL_418425|2020- .................... .................... .................... .................... .................... ....................
A 312 hCoV-19/France/ARA-10251/2020|EPI_ISL_418426|2020- .................... .................... .................... .................... .................... ....................
A 313 hCoV-19/France/ARA-10257/2020|EPI_ISL_418427|2020- .................... .................... .................... .................... .................... ....................
A 314 hCoV-19/France/ARA-10282/2020|EPI_ISL_418428|2020- .................... .................... .................... .................... .................... ....................
A 315 hCoV-19/France/ARA-10876/2020|EPI_ISL_418429|2020- .................... .................... .................... .................... .................... ....................
A 318 hCoV-19/Hangzhou/HZ48/2020|EPI_ISL_418441|2020-01- .................... .................... .................... .................... .................... ....................
```

| | | | | | | | | |
|---|---|---|---|---|---|---|---|---|
| A | 319 hCoV-19/Hangzhou/HZ49/2020\|EPI_ISL_418442\|2020-01- | ···················· | ···················· | ···················· | ···················· | ···················· | ···················· | |
| A | 320 hCoV-19/USA/NY-NYUMC39/2020\|EPI_ISL_419700\|2020-03 | ···················· | ···················· | ···················· | ···················· | ···················· | ···················· | |
| A | 321 hCoV-19/USA/NY-NYUMC41/2020\|EPI_ISL_419702\|2020-03 | ···················· | ···················· | ···················· | ···················· | ···················· | ···················· | |
| A | 322 hCoV-19/USA/NY-NYUMC43/2020\|EPI_ISL_419704\|2020-03 | ···················· | ···················· | ···················· | ···················· | ···················· | ···················· | |
| A | 323 hCoV-19/USA/NY-NYUMC44/2020\|EPI_ISL_419705\|2020-03 | ···················· | ···················· | ···················· | ···················· | ···················· | ···················· | |
| A | 324 hCoV-19/USA/VA-DCLS-0012/2020\|EPI_ISL_419706\|2020- | ···················· | ···················· | ···················· | ···················· | ···················· | ···················· | |
| A | 325 hCoV-19/Australia/VIC31/2020\|EPI_ISL_419750\|2020-0 | ···················· | ···················· | ···················· | ···················· | ···················· | ···················· | |
| A | 326 hCoV-19/Australia/VIC32/2020\|EPI_ISL_419751\|2020-0 | ···················· | ···················· | ···················· | ···················· | ···················· | ···················· | |
| A | 327 hCoV-19/Australia/VIC33/2020\|EPI_ISL_419752\|2020-0 | ···················· | ···················· | ···················· | ···················· | ···················· | ···················· | |
| A | 328 hCoV-19/Australia/VIC34/2020\|EPI_ISL_419753\|2020-0 | ···················· | ···················· | ···················· | ···················· | ···················· | ···················· | |
| A | 329 hCoV-19/Australia/VIC37/2020\|EPI_ISL_419754\|2020-0 | ···················· | ···················· | ···················· | ···················· | ···················· | ···················· | |
| A | 330 hCoV-19/Australia/VIC35/2020\|EPI_ISL_419755\|2020-0 | ···················· | ···················· | ···················· | ···················· | ···················· | ···················· | |
| A | 331 hCoV-19/Australia/VIC38/2020\|EPI_ISL_419756\|2020-0 | ···················· | ···················· | ···················· | ···················· | ···················· | ···················· | |
| A | 332 hCoV-19/Australia/VIC39/2020\|EPI_ISL_419757\|2020-0 | ···················· | ···················· | ···················· | ···················· | ···················· | ···················· | |
| A | 333 hCoV-19/Australia/VIC40/2020\|EPI_ISL_419758\|2020-0 | ···················· | ···················· | ···················· | ···················· | ···················· | ···················· | |
| A | 334 hCoV-19/Australia/VIC47/2020\|EPI_ISL_419759\|2020-0 | ···················· | ···················· | ···················· | ···················· | ···················· | ···················· | |
| A | 335 hCoV-19/Australia/VIC36/2020\|EPI_ISL_419765\|2020-0 | ···················· | ···················· | ···················· | ···················· | ···················· | ···················· | |
| A | 336 hCoV-19/Australia/VIC50/2020\|EPI_ISL_419766\|2020-0 | ···················· | ···················· | ···················· | ···················· | ···················· | ···················· | |
| A | 337 hCoV-19/Australia/VIC44/2020\|EPI_ISL_419767\|2020-0 | ···················· | ···················· | ···················· | ···················· | ···················· | ···················· | |
| A | 338 hCoV-19/Australia/VIC53/2020\|EPI_ISL_419769\|2020-0 | ···················· | ···················· | ···················· | ···················· | ···················· | ···················· | |
| A | 339 hCoV-19/Australia/VIC54/2020\|EPI_ISL_419770\|2020-0 | ···················· | ···················· | ···················· | ···················· | ···················· | ···················· | |
| A | 340 hCoV-19/Australia/VIC59/2020\|EPI_ISL_419771\|2020-0 | ···················· | ···················· | ···················· | ···················· | ···················· | ···················· | |
| A | 341 hCoV-19/Australia/VIC60/2020\|EPI_ISL_419772\|2020-0 | ···················· | ···················· | ···················· | ···················· | ···················· | ···················· | |
| A | 342 hCoV-19/Australia/VIC61/2020\|EPI_ISL_419773\|2020-0 | ···················· | ···················· | ···················· | ···················· | ···················· | ···················· | |
| A | 343 hCoV-19/Australia/VIC62/2020\|EPI_ISL_419774\|2020-0 | ···················· | ···················· | ···················· | ···················· | ···················· | ···················· | |
| A | 344 hCoV-19/Australia/VIC63/2020\|EPI_ISL_419775\|2020-0 | ···················· | ···················· | ···················· | ···················· | ···················· | ···················· | |
| A | 345 hCoV-19/Australia/VIC55/2020\|EPI_ISL_419776\|2020-0 | ···················· | ···················· | ···················· | ···················· | ···················· | ···················· | |
| A | 346 hCoV-19/Australia/VIC56/2020\|EPI_ISL_419777\|2020-0 | ···················· | ···················· | ···················· | ···················· | ···················· | ···················· | |
| A | 347 hCoV-19/Australia/VIC64/2020\|EPI_ISL_419778\|2020-0 | ···················· | ···················· | ···················· | ···················· | ···················· | ···················· | |
| A | 348 hCoV-19/Australia/VIC57/2020\|EPI_ISL_419779\|2020-0 | ···················· | ···················· | ···················· | ···················· | ···················· | ···················· | |
| A | 349 hCoV-19/Australia/VIC66/2020\|EPI_ISL_419780\|2020-0 | ···················· | ···················· | ···················· | ···················· | ···················· | ···················· | |
| A | 350 hCoV-19/Australia/VIC67/2020\|EPI_ISL_419781\|2020-0 | ···················· | ···················· | ···················· | ···················· | ···················· | ···················· | |
| A | 351 hCoV-19/Australia/VIC68/2020\|EPI_ISL_419782\|2020-0 | ···················· | ···················· | ···················· | ···················· | ···················· | ···················· | |
| A | 352 hCoV-19/Australia/VIC69/2020\|EPI_ISL_419783\|2020-0 | ···················· | ···················· | ···················· | ···················· | ···················· | ···················· | |
| A | 353 hCoV-19/Australia/VIC58/2020\|EPI_ISL_419784\|2020-0 | ···················· | ···················· | ···················· | ···················· | ···················· | ···················· | |
| A | 354 hCoV-19/Australia/VIC73/2020\|EPI_ISL_419785\|2020-0 | ···················· | ···················· | ···················· | ···················· | ···················· | ···················· | |
| A | 355 hCoV-19/Australia/VIC70/2020\|EPI_ISL_419786\|2020-0 | ···················· | ···················· | ···················· | ···················· | ···················· | ···················· | |
| A | 356 hCoV-19/Australia/VIC75/2020\|EPI_ISL_419787\|2020-0 | ···················· | ···················· | ···················· | ···················· | ···················· | ···················· | |
| A | 357 hCoV-19/Australia/VIC76/2020\|EPI_ISL_419788\|2020-0 | ···················· | ···················· | ···················· | ···················· | ···················· | ···················· | |
| A | 358 hCoV-19/Australia/VIC77/2020\|EPI_ISL_419789\|2020-0 | ···················· | ···················· | ···················· | ···················· | ···················· | ···················· | |
| A | 360 hCoV-19/England/20139060004/2020\|EPI_ISL_420731\|20 | ···················· | ···················· | ···················· | ···················· | ···················· | ···················· | |
| A | 361 hCoV-19/England/20139060404/2020\|EPI_ISL_420732\|20 | ···················· | ···················· | ···················· | ···················· | ···················· | ···················· | |
| A | 362 hCoV-19/England/20139060504/2020\|EPI_ISL_420733\|20 | ···················· | ···················· | ···················· | ···················· | ···················· | ···················· | |
| A | 363 hCoV-19/England/20139060804/2020\|EPI_ISL_420734\|20 | ···················· | ···················· | ···················· | ···················· | ···················· | ···················· | |
| A | 364 hCoV-19/England/20139061104/2020\|EPI_ISL_420735\|20 | ···················· | ···················· | ···················· | ···················· | ···················· | ···N···N·N········ | 1 |
| A | 365 hCoV-19/England/20139061704/2020\|EPI_ISL_420736\|20 | ···················· | ···················· | ···················· | ···················· | ···················· | ···················· | |
| A | 367 hCoV-19/England/20139062704/2020\|EPI_ISL_420742\|20 | ···················· | ···················· | ···················· | ···················· | ···················· | ···················· | |
| A | 368 hCoV-19/England/20139062804/2020\|EPI_ISL_420743\|20 | ···················· | ···················· | ···················· | ···················· | ···················· | ···················· | |
| A | 370 hCoV-19/England/20139063504/2020\|EPI_ISL_420745\|20 | ···················· | ···················· | ···················· | ···················· | ···················· | ···················· | |
| A | 371 hCoV-19/England/20139063604/2020\|EPI_ISL_420746\|20 | ···················· | ···················· | ···················· | ···················· | ···················· | ···················· | |
| A | 372 hCoV-19/England/20139063704/2020\|EPI_ISL_420747\|20 | ···················· | ···················· | ···················· | ···················· | ···················· | ···················· | |
| A | 373 hCoV-19/England/20139063804/2020\|EPI_ISL_420748\|20 | ···················· | ···················· | ···················· | ···················· | ···················· | ···················· | |
| A | 374 hCoV-19/England/20140037504/2020\|EPI_ISL_420749\|20 | ···················· | ···················· | ···················· | ···················· | ···················· | ···················· | |
| A | 375 hCoV-19/England/20140039004/2020\|EPI_ISL_420752\|20 | ···················· | ···················· | ···················· | ···················· | ···················· | ··N················· | |
| A | 376 hCoV-19/England/20140039104/2020\|EPI_ISL_420753\|20 | ···················· | ···················· | ···················· | ···················· | ·····C·············· | ···················· | |
| A | 377 hCoV-19/England/20140039304/2020\|EPI_ISL_420755\|20 | ···················· | ···················· | ···················· | ···················· | ···················· | ···················· | |
| A | 378 hCoV-19/England/20140039404/2020\|EPI_ISL_420756\|20 | ···················· | ···················· | ···················· | ···················· | ···················· | ···················· | |
| A | 379 hCoV-19/England/20140039604/2020\|EPI_ISL_420758\|20 | ···················· | ···················· | ···················· | ···················· | ·····C·············· | ···················· | |
| A | 380 hCoV-19/England/20140048704/2020\|EPI_ISL_420761\|20 | ···················· | ···················· | ···················· | ···················· | ···················· | ···················· | |
| A | 381 hCoV-19/England/20142022504/2020\|EPI_ISL_420765\|20 | ···················· | ···················· | ···················· | ···················· | ···················· | ···················· | |
| A | 382 hCoV-19/England/20142060404/2020\|EPI_ISL_420766\|20 | ···················· | ···················· | ···················· | ···················· | ···················· | ···················· | |
| A | 383 hCoV-19/England/20142060804/2020\|EPI_ISL_420767\|20 | ···················· | ···················· | ···················· | ···················· | ···················· | ···················· | |
| A | 384 hCoV-19/England/20142061004/2020\|EPI_ISL_420769\|20 | ···················· | ···················· | ···················· | ···················· | ···················· | ···················· | |
| A | 385 hCoV-19/England/20142061804/2020\|EPI_ISL_420772\|20 | ···················· | ···················· | ···················· | ···················· | ···················· | ···················· | |
| A | 387 hCoV-19/USA/WA-UW-1682/2020\|EPI_ISL_424218\|2020-03 | ···················· | ···················· | ···················· | ···················· | ···················· | ···················· | |
| A | 388 hCoV-19/USA/WA-UW-1695/2020\|EPI_ISL_424219\|2020-03 | ···················· | ···················· | ···················· | ···················· | ···················· | ···················· | |
| A | 390 hCoV-19/USA/WA-UW-1748/2020\|EPI_ISL_424221\|2020-03 | ···················· | ···················· | ···················· | ···················· | ···················· | ···················· | |
| A | 391 hCoV-19/USA/WA-UW-1700/2020\|EPI_ISL_424222\|2020-03 | ···················· | ···················· | ···················· | ···················· | ···················· | ···················· | |
| A | 392 hCoV-19/USA/WA-UW-1744/2020\|EPI_ISL_424223\|2020-03 | ···················· | ···················· | ···················· | ···················· | ···················· | ···················· | |
| A | 393 hCoV-19/USA/WA-UW-1745/2020\|EPI_ISL_424224\|2020-03 | ···················· | ···················· | ···················· | ···················· | ···················· | ···················· | |
| A | 394 hCoV-19/USA/WA-UW-1707/2020\|EPI_ISL_424225\|2020-03 | ···················· | ···················· | ···················· | ···················· | ···················· | ···················· | |
| A | 395 hCoV-19/USA/WA-UW-1705/2020\|EPI_ISL_424226\|2020-03 | ···················· | ···················· | ···················· | ···················· | ···················· | ···················· | |
| A | 396 hCoV-19/USA/WA-UW-1706/2020\|EPI_ISL_424227\|2020-03 | ···················· | ···················· | ···················· | ···················· | ···················· | ···················· | |
| A | 397 hCoV-19/USA/WA-UW-1702/2020\|EPI_ISL_424228\|2020-03 | ···················· | ···················· | ···················· | ···················· | ···················· | ···················· | |
| A | 398 hCoV-19/USA/WA-UW-1724/2020\|EPI_ISL_424229\|2020-03 | ···················· | ···················· | ···················· | ···················· | ···················· | ···················· | |
| A | 399 hCoV-19/USA/WA-UW-1709/2020\|EPI_ISL_424230\|2020-03 | ···················· | ···················· | ···················· | ···················· | ···················· | ···················· | |
| A | 400 hCoV-19/USA/WA-UW-1731/2020\|EPI_ISL_424231\|2020-03 | ···················· | ···················· | ···················· | ···················· | ···················· | ···················· | |
| A | 401 hCoV-19/USA/WA-UW-1708/2020\|EPI_ISL_424232\|2020-03 | ···················· | ···················· | ···················· | ···················· | ···················· | ···················· | |

| | | | | | | | | |
|---|---|---|---|---|---|---|---|---|
| A | 402 | hCoV-19/USA/WA-UW-1772/2020\|EPI_ISL_424233\|2020-03 | ···················· | ···················· | ···················· | ···················· | ···················· | ···················· |
| A | 403 | hCoV-19/USA/WA-UW-1722/2020\|EPI_ISL_424234\|2020-03 | ···················· | ···················· | ···················· | ···················· | ···················· | ···················· |
| A | 404 | hCoV-19/USA/WA-UW-1733/2020\|EPI_ISL_424235\|2020-03 | ···················· | ···················· | ···················· | ···················· | ···················· | ···················· |
| A | 405 | hCoV-19/USA/WA-UW-1732/2020\|EPI_ISL_424236\|2020-03 | ···················· | ···················· | ···················· | ···················· | ···················· | ···················· |
| A | 406 | hCoV-19/USA/WA-UW-1739/2020\|EPI_ISL_424237\|2020-03 | ···················· | ···················· | ···················· | ···················· | ···················· | ···················· |
| A | 407 | hCoV-19/USA/WA-UW-1741/2020\|EPI_ISL_424238\|2020-03 | ···················· | ···················· | ···················· | ···················· | ···················· | ···················· |
| A | 408 | hCoV-19/USA/WA-UW-1750/2020\|EPI_ISL_424239\|2020-03 | ···················· | ···················· | ···················· | ···················· | ···················· | ···················· |
| A | 409 | hCoV-19/USA/WA-UW-1730/2020\|EPI_ISL_424240\|2020-03 | ···················· | ···················· | ···················· | ···················· | ···················· | ···················· |
| A | 410 | hCoV-19/USA/WA-UW-1735/2020\|EPI_ISL_424241\|2020-03 | ···················· | ···················· | ···················· | ···················· | ···················· | ···················· |
| A | 411 | hCoV-19/USA/WA-UW-1753/2020\|EPI_ISL_424242\|2020-03 | ···················· | ···················· | ···················· | ···················· | ···················· | ···················· |
| A | 412 | hCoV-19/USA/WA-UW-1756/2020\|EPI_ISL_424243\|2020-03 | ···················· | ···················· | ···················· | ···················· | ···················· | ···················· |
| A | 413 | hCoV-19/USA/WA-UW-1770/2020\|EPI_ISL_424244\|2020-03 | ···················· | ···················· | ···················· | ···················· | ···················· | ···················· |
| A | 414 | hCoV-19/USA/WA-UW-1762/2020\|EPI_ISL_424245\|2020-03 | ···················· | ···················· | ···················· | ···················· | ···················· | ···················· |
| A | 415 | hCoV-19/USA/WA-UW-1758/2020\|EPI_ISL_424246\|2020-03 | ···················· | ···················· | ···················· | ···················· | ···················· | ···················· |
| A | 416 | hCoV-19/USA/WA-UW-1760/2020\|EPI_ISL_424247\|2020-03 | ···················· | ···················· | ···················· | ···················· | ···················· | ···················· |
| A | 417 | hCoV-19/USA/UN-UW-1773/2020\|EPI_ISL_424248\|2020-03 | ···················· | ···················· | ···················· | ···················· | ···················· | ···················· |
| A | 418 | hCoV-19/USA/UN-UW-1791/2020\|EPI_ISL_424249\|2020-03 | ···················· | ···················· | ···················· | ···················· | ···················· | ···················· |
| A | 419 | hCoV-19/USA/WA-UW-1785/2020\|EPI_ISL_424250\|2020-03 | ···················· | ···················· | ···················· | ···················· | ···················· | ···················· |
| A | 420 | hCoV-19/USA/WA-UW-1801/2020\|EPI_ISL_424251\|2020-03 | ···················· | ···················· | ···················· | ···················· | ···················· | ···················· |
| A | 421 | hCoV-19/USA/WA-UW-1840/2020\|EPI_ISL_424252\|2020-03 | ···················· | ···················· | ···················· | ···················· | ···················· | ···················· |
| A | 422 | hCoV-19/USA/WA-UW-1792/2020\|EPI_ISL_424253\|2020-03 | ···················· | ···················· | ···················· | ···················· | ···················· | ···················· |
| A | 423 | hCoV-19/USA/WA-UW-1796/2020\|EPI_ISL_424254\|2020-03 | ···················· | ···················· | ···················· | ···················· | ···················· | ···················· |
| A | 424 | hCoV-19/USA/UN-UW-1832/2020\|EPI_ISL_424255\|2020-03 | ···················· | ···················· | ···················· | ···················· | ···················· | ···················· |
| A | 425 | hCoV-19/USA/WA-UW-1774/2020\|EPI_ISL_424256\|2020-03 | ···················· | ···················· | ···················· | ···················· | ···················· | ···················· |
| A | 426 | hCoV-19/USA/WA-UW-1775/2020\|EPI_ISL_424257\|2020-03 | ···················· | ···················· | ···················· | ···················· | ···················· | ···················· |
| A | 427 | hCoV-19/USA/WA-UW-1778/2020\|EPI_ISL_424258\|2020-03 | ···················· | ···················· | ···················· | ···················· | ···················· | ···················· |
| A | 428 | hCoV-19/USA/WA-UW-1797/2020\|EPI_ISL_424259\|2020-03 | ···················· | ···················· | ···················· | ···················· | ···················· | ···················· |
| A | 429 | hCoV-19/USA/WA-UW-1779/2020\|EPI_ISL_424260\|2020-03 | ···················· | ···················· | ···················· | ···················· | ···················· | ···················· |
| A | 430 | hCoV-19/USA/WA-UW-1784/2020\|EPI_ISL_424261\|2020-03 | ···················· | ···················· | ···················· | ···················· | ···················· | ···················· |
| A | 431 | hCoV-19/USA/WA-UW-1782/2020\|EPI_ISL_424262\|2020-03 | ···················· | ···················· | ···················· | ···················· | ···················· | ···················· |
| A | 432 | hCoV-19/USA/WA-UW-1799/2020\|EPI_ISL_424263\|2020-03 | ···················· | ···················· | ···················· | ···················· | ···················· | ···················· |
| A | 433 | hCoV-19/USA/WA-UW-1805/2020\|EPI_ISL_424264\|2020-03 | ···················· | ···················· | ···················· | ···················· | ···················· | ···················· |
| A | 434 | hCoV-19/USA/UN-UW-1818/2020\|EPI_ISL_424266\|2020-03 | ···················· | ···················· | ···················· | ···················· | ···················· | ···················· |
| A | 435 | hCoV-19/USA/UN-UW-1817/2020\|EPI_ISL_424267\|2020-03 | ···················· | ···················· | ···················· | ···················· | ···················· | ···················· |
| A | 436 | hCoV-19/USA/UN-UW-1816/2020\|EPI_ISL_424268\|2020-03 | ···················· | ···················· | ···················· | ···················· | ···················· | ···················· |
| A | 437 | hCoV-19/USA/WA-UW-1828/2020\|EPI_ISL_424269\|2020-03 | ···················· | ···················· | ···················· | ···················· | ···················· | ···················· |
| A | 438 | hCoV-19/USA/WA-UW-1831/2020\|EPI_ISL_424270\|2020-03 | ···················· | ···················· | ···················· | ···················· | ···················· | ···················· |
| A | 439 | hCoV-19/USA/WA-UW-1827/2020\|EPI_ISL_424271\|2020-03 | ···················· | ···················· | ···················· | ···················· | ···················· | ···················· |
| A | 440 | hCoV-19/USA/WA-UW-1826/2020\|EPI_ISL_424273\|2020-03 | ···················· | ···················· | ···················· | ···················· | ···················· | ···················· |
| A | 441 | hCoV-19/USA/WA-UW-1821/2020\|EPI_ISL_424275\|2020-03 | ···················· | ···················· | ···················· | ···················· | ···················· | ···················· |
| A | 442 | hCoV-19/USA/WA-UW-1822/2020\|EPI_ISL_424276\|2020-03 | ···················· | ···················· | ···················· | ···················· | ···················· | ···················· |
| A | 443 | hCoV-19/USA/WA-UW-1866/2020\|EPI_ISL_424278\|2020-03 | ···················· | ···················· | ···················· | ···················· | ···················· | ···················· |
| A | 444 | hCoV-19/USA/WA-UW-1850/2020\|EPI_ISL_424279\|2020-03 | ···················· | ···················· | ···················· | ···················· | ···················· | ···················· |
| A | 445 | hCoV-19/USA/WA-UW-1853/2020\|EPI_ISL_424290\|2020-03 | ···················· | ···················· | ···················· | ···················· | ···················· | ···················· |
| A | 446 | hCoV-19/USA/WA-UW-1854/2020\|EPI_ISL_424291\|2020-03 | ···················· | ···················· | ···················· | ···················· | ···················· | ···················· |
| A | 447 | hCoV-19/USA/WA-UW-1855/2020\|EPI_ISL_424292\|2020-03 | ···················· | ···················· | ···················· | ···················· | ···················· | ···················· |
| A | 448 | hCoV-19/USA/WA-UW-1870/2020\|EPI_ISL_424293\|2020-03 | ···················· | ···················· | ···················· | ···················· | ···················· | ···················· |
| A | 449 | hCoV-19/USA/WA-UW-1872/2020\|EPI_ISL_424294\|2020-03 | ···················· | ···················· | ···················· | ···················· | ···················· | ···················· |
| A | 450 | hCoV-19/USA/WA-UW-1905/2020\|EPI_ISL_424295\|2020-03 | ···················· | ···················· | ···················· | ···················· | ···················· | ···················· |
| A | 451 | hCoV-19/USA/WA-UW-1919/2020\|EPI_ISL_424296\|2020-03 | ···················· | ···················· | ···················· | ···················· | ···················· | ···················· |
| A | 452 | hCoV-19/USA/WA-UW-1913/2020\|EPI_ISL_424297\|2020-03 | ···················· | ···················· | ···················· | ···················· | ···················· | ···················· |
| A | 453 | hCoV-19/USA/WA-UW-1920/2020\|EPI_ISL_424298\|2020-03 | ···················· | ···················· | ···················· | ···················· | ···················· | ···················· |
| A | 454 | hCoV-19/USA/WA-UW-1904/2020\|EPI_ISL_424299\|2020-03 | ···················· | ···················· | ···················· | ···················· | ···················· | ···················· |
| A | 455 | hCoV-19/England/NOTT-10E46F/2020\|EPI_ISL_425548\|20 | ···················· | ···················· | ···················· | ···················· | ···················· | ···················· |
| A | 456 | hCoV-19/England/NOTT-10E47E/2020\|EPI_ISL_425549\|20 | ···················· | ···················· | ···················· | ···················· | ···················· | ···················· |
| A | 457 | hCoV-19/England/NOTT-10E48D/2020\|EPI_ISL_425550\|20 | ···················· | ···················· | ···················· | ···················· | ···················· | ···················· |
| A | 458 | hCoV-19/England/NOTT-10E49C/2020\|EPI_ISL_425551\|20 | ···················· | ···················· | ···················· | ···················· | ···················· | ···················· |
| A | 459 | hCoV-19/England/NOTT-10E4C9/2020\|EPI_ISL_425554\|20 | ···················· | ···················· | ···················· | ···················· | ···················· | ···················· |
| A | 460 | hCoV-19/England/NOTT-10E55D/2020\|EPI_ISL_425561\|20 | ···················· | ···················· | ···················· | ···················· | ···················· | ···················· |
| A | 461 | hCoV-19/England/NOTT-10E57B/2020\|EPI_ISL_425562\|20 | ···················· | ···················· | ···················· | ···················· | ···················· | ···················· |
| A | 462 | hCoV-19/England/NOTT-10E58A/2020\|EPI_ISL_425563\|20 | ···················· | ···················· | ···················· | ···················· | ···················· | ···················· |
| A | 463 | hCoV-19/England/NOTT-10E5A8/2020\|EPI_ISL_425565\|20 | ···················· | ···················· | ···················· | ···················· | ···················· | ···················· |
| A | 464 | hCoV-19/England/NOTT-10E5B7/2020\|EPI_ISL_425566\|20 | ···················· | ···················· | ···················· | ···················· | ···················· | ···················· |
| A | 465 | hCoV-19/England/NOTT-10E5C6/2020\|EPI_ISL_425567\|20 | ···················· | ···················· | ···················· | ···················· | ···················· | ···················· |
| A | 466 | hCoV-19/England/NOTT-10E5D5/2020\|EPI_ISL_425568\|20 | ···················· | ···················· | ···················· | ···················· | ···················· | ···················· |
| A | 467 | hCoV-19/England/NOTT-10E5F3/2020\|EPI_ISL_425570\|20 | ···················· | ···················· | ···················· | ···················· | ···················· | ···················· |
| A | 468 | hCoV-19/England/NOTT-10E61E/2020\|EPI_ISL_425571\|20 | ···················· | ···················· | ···················· | ···················· | ···················· | ···················· |
| A | 469 | hCoV-19/England/NOTT-10E64B/2020\|EPI_ISL_425573\|20 | ···················· | ···················· | ···················· | ···················· | ···················· | ···················· |
| A | 470 | hCoV-19/England/NOTT-10E65A/2020\|EPI_ISL_425574\|20 | ···················· | ···················· | ···················· | ···················· | ···················· | ···················· |
| A | 471 | hCoV-19/England/NOTT-10E669/2020\|EPI_ISL_425575\|20 | ···················· | ···················· | ···················· | ···················· | ···················· | ···················· |
| A | 472 | hCoV-19/England/NOTT-10E678/2020\|EPI_ISL_425576\|20 | ···················· | ···················· | ···················· | ···················· | ···················· | ···················· |
| A | 473 | hCoV-19/England/NOTT-10E6A5/2020\|EPI_ISL_425579\|20 | ···················· | ···················· | ···················· | ···················· | ···················· | ···················· |
| A | 474 | hCoV-19/England/NOTT-10E6C3/2020\|EPI_ISL_425581\|20 | ···················· | ···················· | ···················· | ···················· | ···················· | ···················· |
| A | 475 | hCoV-19/England/NOTT-10E6D2/2020\|EPI_ISL_425582\|20 | ···················· | ···················· | ···················· | ···················· | ···················· | ···················· |
| A | 476 | hCoV-19/England/NOTT-10E70C/2020\|EPI_ISL_425583\|20 | ···················· | ···················· | ···················· | ···················· | ···················· | ···················· |
| A | 477 | hCoV-19/England/NOTT-10E71B/2020\|EPI_ISL_425584\|20 | ···················· | ···················· | ···················· | ···················· | ···················· | ···················· |
| A | 478 | hCoV-19/England/NOTT-10E739/2020\|EPI_ISL_425586\|20 | ···················· | ···················· | ···················· | ···················· | ···················· | ···················· |
| A | 479 | hCoV-19/England/NOTT-10E748/2020\|EPI_ISL_425587\|20 | ···················· | ···················· | ···················· | ···················· | ···················· | ···················· |

| | | | | | | | | | |
|---|---|---|---|---|---|---|---|---|---|
| A | 480 | hCoV-19/England/NOTT-10E775/2020\|EPI_ISL_425590\|20 | ···················· | ···················· | ···················· | ···················· | ···················· | ···················· | |
| A | 481 | hCoV-19/England/NOTT-10E7A2/2020\|EPI_ISL_425593\|20 | ···················· | ···················· | ···················· | ···················· | ···················· | ···················· | |
| A | 482 | hCoV-19/Australia/VIC912/2020\|EPI_ISL_426881\|2020- | ···················· | ···················· | ···················· | ···················· | ···················· | ···················· | |
| A | 484 | hCoV-19/Czech Republic/IAB_1/2020\|EPI_ISL_426883\|2 | ···················· | ···················· | ···················· | ···················· | ···················· | ···················· | |
| A | 485 | hCoV-19/Czech Republic/IAB_4/2020\|EPI_ISL_426884\|2 | ···················· | ···················· | ···················· | ···················· | ···················· | ···················· | |
| A | 486 | hCoV-19/Czech Republic/IAB_8/2020\|EPI_ISL_426885\|2 | ···················· | ···················· | ···················· | ···················· | ···················· | ···················· | |
| A | 487 | hCoV-19/Czech Republic/IAB_9/2020\|EPI_ISL_426886\|2 | ···················· | ···················· | ···················· | ···················· | ···················· | ···················· | |
| A | 488 | hCoV-19/Czech Republic/IAB_10/2020\|EPI_ISL_426887\| | ···················· | ···················· | ···················· | ···················· | ···················· | ···················· | |
| A | 489 | hCoV-19/Czech Republic/IAB_12/2020\|EPI_ISL_426888\| | ···················· | ···················· | ···················· | ···················· | ···················· | ···················· | |
| A | 490 | hCoV-19/Czech Republic/IAB_14/2020\|EPI_ISL_426889\| | ···················· | ···················· | ···················· | ···················· | ···················· | ···············A···· | |
| A | 491 | hCoV-19/Czech Republic/IAB_15/2020\|EPI_ISL_426890\| | ···················· | ···················· | ···················· | ···················· | ···················· | ···················· | |
| A | 492 | hCoV-19/Czech Republic/IAB_16/2020\|EPI_ISL_426891\| | ···················· | ···················· | ···················· | ···················· | ···················· | ···················· | |
| A | 493 | hCoV-19/Czech Republic/IAB_17/2020\|EPI_ISL_426892\| | ···················· | ···················· | ···················· | ···················· | ···················· | ···················· | |
| A | 494 | hCoV-19/Czech Republic/IAB_18/2020\|EPI_ISL_426893\| | ···················· | ···················· | ···················· | ···················· | ···················· | ···················· | |
| A | 495 | hCoV-19/Czech Republic/IAB_20/2020\|EPI_ISL_426894\| | ···················· | ···················· | ···················· | ···················· | ···················· | ···················· | |
| A | 496 | hCoV-19/Czech Republic/IAB_21/2020\|EPI_ISL_426895\| | ···················· | ···················· | ···················· | ···················· | ···················· | ···················· | |
| A | 497 | hCoV-19/Czech Republic/IAB_22/2020\|EPI_ISL_426896\| | ···················· | ···················· | ···················· | ···················· | ···················· | ···················· | |
| A | 498 | hCoV-19/Czech Republic/IAB_23/2020\|EPI_ISL_426897\| | ···················· | ···················· | ···················· | ···················· | ···················· | ···················· | |
| A | 501 | hCoV-19/Guangzhou/GZMU0057/2020\|EPI_ISL_429100\|202 | ···················· | ···················· | ···················· | ···················· | ···················· | ···················· | |
| A | 502 | hCoV-19/Guangzhou/GZMU0058/2020\|EPI_ISL_429101\|202 | ···················· | ···················· | ···················· | ···················· | ···················· | ···················· | |
| A | 503 | hCoV-19/Guangzhou/GZMU0059/2020\|EPI_ISL_429102\|202 | ···················· | ···················· | ···················· | ···················· | ···················· | ···················· | |
| A | 504 | hCoV-19/Guangzhou/GZMU0060/2020\|EPI_ISL_429103\|202 | ···················· | ···················· | ···················· | ···················· | ···················· | ···················· | |
| A | 505 | hCoV-19/Guangzhou/GZMU0036/2020\|EPI_ISL_429104\|202 | ···················· | ···················· | ···················· | ···················· | ···················· | ···················· | |
| A | 506 | hCoV-19/Guangzhou/GZMU0055/2020\|EPI_ISL_429105\|202 | ···················· | ···················· | ···················· | ···················· | ···················· | ···················· | |
| A | 507 | hCoV-19/Wuhan/OS52/2020\|EPI_ISL_450500\|2020-01-10 | ···················· | ···················· | ···················· | ···················· | ···················· | ···················· | |
| A | 508 | hCoV-19/Wuhan/YB012611/2020\|EPI_ISL_450501\|2020-01 | ···················· | ···················· | ···················· | ···················· | ···················· | ···················· | |
| A | 509 | hCoV-19/Wuhan/YB012605/2020\|EPI_ISL_450502\|2020-01 | ···················· | ···················· | ···················· | ···················· | ···················· | ···················· | |
| A | 510 | hCoV-19/Wuhan/YB012602/2020\|EPI_ISL_450503\|2020-01 | ···················· | ···················· | ···················· | ···················· | ···················· | ···················· | |
| A | 511 | hCoV-19/Wuhan/YB012506/2020\|EPI_ISL_450504\|2020-01 | ···················· | ···················· | ···················· | ···················· | ···················· | ···················· | |
| A | 512 | hCoV-19/Brazil/SP02cc/2020\|EPI_ISL_450506\|2020-03- | ···················· | ···················· | ···················· | ···················· | ···················· | ···················· | |
| A | 513 | hCoV-19/South Korea/KUMC03/2020\|EPI_ISL_413513\|202 | ···················· | ···················· | ···················· | ···················· | ···················· | ···················· | |
| A | 514 | hCoV-19/South Korea/KUMC04/2020\|EPI_ISL_413514\|202 | ···················· | ···················· | ···················· | ···················· | ···················· | ···················· | |
| A | 515 | hCoV-19/South Korea/KUMC05/2020\|EPI_ISL_413515\|202 | ···················· | ···················· | ···················· | ···················· | ···················· | ···················· | |
| A | 516 | hCoV-19/South Korea/KUMC06/2020\|EPI_ISL_413516\|202 | ···················· | ···················· | ···················· | ···················· | ···················· | ···················· | |
| A | 517 | hCoV-19/Beijing/105/2020\|EPI_ISL_413518\|2020-01-26 | ···················· | ···················· | ···················· | ···················· | ···················· | ···················· | |
| A | 518 | hCoV-19/Beijing/231/2020\|EPI_ISL_413519\|2020-01-28 | ···················· | ···················· | ···················· | ···················· | ···················· | ···················· | |
| A | 519 | hCoV-19/Beijing/233/2020\|EPI_ISL_413520\|2020-01-28 | ···················· | ···················· | ···················· | ···················· | ···················· | ···················· | |
| A | 520 | hCoV-19/Beijing/235/2020\|EPI_ISL_413521\|2020-01-28 | ···················· | ···················· | ···················· | ···················· | ···················· | ···················· | |
| A | 521 | hCoV-19/India/MH-1-27/2020\|EPI_ISL_413522\|2020-01- | ···················· | ···················· | ···················· | ···················· | ···················· | ···················· | |
| A | 522 | hCoV-19/India/MH-1-31/2020\|EPI_ISL_413523\|2020-01- | ···················· | ···················· | ···················· | ···················· | ···················· | ···················· | |
| A | 523 | hCoV-19/Wales/PHW1/2020\|EPI_ISL_413555\|2020-02-27 | ···················· | ···················· | ···················· | ···················· | ···················· | ···················· | |
| A | 524 | hCoV-19/Wales/PHW2/2020\|EPI_ISL_413556\|2020-03-04 | ···················· | ···················· | ···················· | ···················· | ···················· | ···················· | |
| A | 525 | hCoV-19/USA/CA-CDPH-UC1/2020\|EPI_ISL_413557\|2020-0 | ···················· | ···················· | ···················· | ···················· | ···················· | ···················· | |
| A | 526 | hCoV-19/USA/CA-CDPH-UC2/2020\|EPI_ISL_413558\|2020-0 | ···················· | ···················· | ···················· | ···················· | ···················· | ···················· | |
| A | 527 | hCoV-19/USA/CA-CDPH-UC3/2020\|EPI_ISL_413559\|2020-0 | ···················· | ···················· | ···················· | ···················· | ···················· | ···················· | |
| A | 528 | hCoV-19/USA/WA-S3/2020\|EPI_ISL_413560\|2020-02-28 | ···················· | ···················· | ···················· | ···················· | ···················· | ···················· | |
| A | 529 | hCoV-19/USA/CA-CDPH-UC4/2020\|EPI_ISL_413561\|2020-0 | ···················· | ···················· | ···················· | ···················· | ···················· | ···················· | |
| A | 530 | hCoV-19/USA/WA11-UW7/2020\|EPI_ISL_413562\|2020-03-0 | ···················· | ···················· | ···················· | ···················· | ···················· | ···················· | |
| A | 531 | hCoV-19/USA/WA12-UW8/2020\|EPI_ISL_413563\|2020-03-0 | ···················· | ···················· | ···················· | ···················· | ···················· | ···················· | |
| A | 532 | hCoV-19/Netherlands/Andel_1365066/2020\|EPI_ISL_413 | ···················· | ···················· | ···················· | ···················· | ···················· | ···················· | |
| A | 533 | hCoV-19/Netherlands/Blaricum_1364780/2020\|EPI_ISL_ | ···················· | ···················· | ···················· | ···················· | ···················· | ···················· | |
| A | 534 | hCoV-19/Netherlands/Dalen_1363624/2020\|EPI_ISL_413 | ···················· | ···················· | ···················· | ···················· | ···················· | ···················· | |
| A | 535 | hCoV-19/USA/WA-S37/2020\|EPI_ISL_417090\|2020-03-04 | ···················· | ···················· | ···················· | ···················· | ···················· | ···················· | |
| A | 536 | hCoV-19/USA/WA-S38/2020\|EPI_ISL_417091\|2020-03-04 | ···················· | ···················· | ···················· | ···················· | ···················· | ···················· | |
| A | 537 | hCoV-19/USA/WA-S39/2020\|EPI_ISL_417092\|2020-03-04 | ···················· | ···················· | ···················· | ···················· | ···················· | ···················· | |
| A | 538 | hCoV-19/USA/WA-S40/2020\|EPI_ISL_417093\|2020-02-28 | ···················· | ···················· | ···················· | ···················· | ···················· | ···················· | |
| A | 539 | hCoV-19/USA/WA-S42/2020\|EPI_ISL_417095\|2020-02-28 | ···················· | ···················· | ···················· | ···················· | ···················· | ···················· | |
| A | 540 | hCoV-19/USA/WA-S43/2020\|EPI_ISL_417096\|2020-02-27 | ···················· | ···················· | ···················· | ···················· | ···················· | ···················· | |
| A | 541 | hCoV-19/USA/WA-S44/2020\|EPI_ISL_417097\|2020-02-28 | ···················· | ···················· | ···················· | ···················· | ···················· | ···················· | |
| A | 542 | hCoV-19/USA/WA-S45/2020\|EPI_ISL_417098\|2020-02-29 | ···················· | ···················· | ···················· | ···················· | ···················· | ···················· | |
| A | 543 | hCoV-19/USA/WA-S46/2020\|EPI_ISL_417099\|2020-02-29 | ···················· | ···················· | ···················· | ···················· | ···················· | ···················· | |
| A | 544 | hCoV-19/England/20108004702/2020\|EPI_ISL_417250\|20 | ···················· | ···················· | ···················· | ···················· | ···················· | ···················· | |
| A | 545 | hCoV-19/England/20108006003/2020\|EPI_ISL_417252\|20 | ···················· | ···················· | ···················· | ···················· | ···················· | ···················· | |
| A | 547 | hCoV-19/England/20108007002/2020\|EPI_ISL_417255\|20 | ···················· | ···················· | ···················· | ···················· | ···················· | ···················· | |
| A | 548 | hCoV-19/England/20108007302/2020\|EPI_ISL_417256\|20 | ···················· | ···················· | ···················· | ···················· | ···················· | ···················· | |
| A | 550 | hCoV-19/England/20108034006/2020\|EPI_ISL_417258\|20 | ···················· | ···················· | ···················· | ···················· | ···················· | ···················· | |
| A | 551 | hCoV-19/England/20109035906/2020\|EPI_ISL_417260\|20 | ···················· | ···················· | ···················· | ···················· | ···················· | ···················· | |
| A | 552 | hCoV-19/England/20109039306/2020\|EPI_ISL_417262\|20 | ···················· | ···················· | ···················· | ···················· | ···················· | ···················· | |
| A | 553 | hCoV-19/England/20109050106/2020\|EPI_ISL_417263\|20 | ···················· | ···················· | ···················· | ···················· | ···················· | ···················· | |
| A | 554 | hCoV-19/England/20109050306/2020\|EPI_ISL_417264\|20 | ···················· | ···················· | ···················· | ···················· | ···················· | ···················· | |
| A | 555 | hCoV-19/England/20109050406/2020\|EPI_ISL_417265\|20 | ···················· | ···················· | ···················· | ···················· | ···················· | ···················· | |
| A | 556 | hCoV-19/England/20109050506/2020\|EPI_ISL_417266\|20 | ···················· | ···················· | ···················· | ···················· | ···················· | ···················· | |
| A | 557 | hCoV-19/England/20109050606/2020\|EPI_ISL_417267\|20 | ···················· | ···················· | ···················· | ···················· | ···················· | ·NNN················ | 1 |
| A | 558 | hCoV-19/England/20109050706/2020\|EPI_ISL_417268\|20 | ···················· | ···················· | ···················· | ···················· | ···················· | ···················· | |
| A | 559 | hCoV-19/England/20109050806/2020\|EPI_ISL_417269\|20 | ···················· | ···················· | ···················· | ···················· | ···················· | ···················· | |
| A | 560 | hCoV-19/England/20109056906/2020\|EPI_ISL_417282\|20 | ···················· | ···················· | ···················· | ···················· | ···················· | ···················· | |
| A | 561 | hCoV-19/England/20109058906/2020\|EPI_ISL_417283\|20 | ···················· | ···················· | ···················· | ···················· | ···················· | ···················· | |
| A | 562 | hCoV-19/England/20109093606/2020\|EPI_ISL_417285\|20 | ···················· | ···················· | ···················· | ···················· | ···················· | ···················· | |

| | | | | | | | | |
|---|---|---|---|---|---|---|---|---|
| A | 563 | hCoV-19/England/20109093706/2020\|EPI_ISL_417286\|20 | ···················· | ···················· | ···················· | ···················· | ···················· | ···················· |
| A | 564 | hCoV-19/England/20109093806/2020\|EPI_ISL_417287\|20 | ···················· | ···················· | ···················· | ···················· | ···················· | ···················· |
| A | 565 | hCoV-19/England/20109093906/2020\|EPI_ISL_417288\|20 | ···················· | ···················· | ···················· | ···················· | ···················· | ···················· |
| A | 566 | hCoV-19/England/20109094006/2020\|EPI_ISL_417289\|20 | ···················· | ···················· | ···················· | ···················· | ···················· | ···················· |
| A | 567 | hCoV-19/Ireland/CO-NVRL-22361/2020\|EPI_ISL_418580\| | ···················· | ···················· | ···················· | ···················· | ···················· | ···················· |
| A | 568 | hCoV-19/Ireland/D-NVRL-22428/2020\|EPI_ISL_418581\|2 | ···················· | ···················· | ···················· | ···················· | ···················· | ···················· |
| A | 569 | hCoV-19/Ireland/LH-NVRL-22901/2020\|EPI_ISL_418582\| | ···················· | ···················· | ···················· | ···················· | ···················· | ···················· |
| A | 570 | hCoV-19/Ireland/D-NVRL-24042/2020\|EPI_ISL_418583\|2 | ···················· | ···················· | ···················· | ···················· | ···················· | ···················· |
| A | 571 | hCoV-19/Ireland/WW-NVRL-24052/2020\|EPI_ISL_418584\| | ···················· | ···················· | ···················· | ···················· | ···················· | ···················· |
| A | 572 | hCoV-19/USA/NY-NYUMC42/2020\|EPI_ISL_419703\|2020-03 | ···················· | ···················· | ···················· | ···················· | ···················· | ···················· |
| A | 574 | hCoV-19/Germany/BY-MVP-V2010492/2020\|EPI_ISL_42089 | ···················· | ···················· | ···················· | ···················· | ···················· | ···················· |
| A | 575 | hCoV-19/USA/WA-UW-1723/2020\|EPI_ISL_424200\|2020-03 | ···················· | ···················· | ···················· | ···················· | ···················· | ···················· |
| A | 576 | hCoV-19/USA/WA-UW-1651/2020\|EPI_ISL_424202\|2020-03 | ···················· | ···················· | ···················· | ···················· | ···················· | ···················· |
| A | 577 | hCoV-19/USA/WA-UW-1652/2020\|EPI_ISL_424203\|2020-03 | ···················· | ···················· | ···················· | ···················· | ···················· | ···················· |
| A | 578 | hCoV-19/USA/WA-UW-1698/2020\|EPI_ISL_424204\|2020-03 | ···················· | ···················· | ···················· | ···················· | ···················· | ···················· |
| A | 579 | hCoV-19/USA/WA-UW-1697/2020\|EPI_ISL_424205\|2020-03 | ···················· | ···················· | ···················· | ···················· | ···················· | ···················· |
| A | 580 | hCoV-19/USA/WA-UW-1716/2020\|EPI_ISL_424207\|2020-03 | ···················· | ···················· | ···················· | ···················· | ···················· | ···················· |
| A | 581 | hCoV-19/USA/WA-UW-1712/2020\|EPI_ISL_424208\|2020-03 | ···················· | ···················· | ···················· | ···················· | ···················· | ···················· |
| A | 582 | hCoV-19/USA/WA-UW-1765/2020\|EPI_ISL_424209\|2020-03 | ···················· | ···················· | ···················· | ···················· | ···················· | ···················· |
| A | 583 | hCoV-19/USA/WA-UW-1718/2020\|EPI_ISL_424210\|2020-03 | ···················· | ···················· | ···················· | ···················· | ···················· | ···················· |
| A | 584 | hCoV-19/USA/WA-UW-1689/2020\|EPI_ISL_424211\|2020-03 | ···················· | ···················· | ···················· | ···················· | ···················· | ···················· |
| A | 585 | hCoV-19/USA/WA-UW-1715/2020\|EPI_ISL_424212\|2020-03 | ···················· | ···················· | ···················· | ···················· | ···················· | ···················· |
| A | 586 | hCoV-19/USA/WA-UW-1670/2020\|EPI_ISL_424213\|2020-03 | ···················· | ···················· | ···················· | ···················· | ···················· | ···················· |
| A | 587 | hCoV-19/USA/WA-UW-1677/2020\|EPI_ISL_424214\|2020-03 | ···················· | ···················· | ···················· | ···················· | ···················· | ···················· |
| A | 588 | hCoV-19/USA/WA-UW-1680/2020\|EPI_ISL_424215\|2020-03 | ···················· | ···················· | ···················· | ···················· | ···················· | ···················· |
| A | 589 | hCoV-19/USA/WA-UW-1690/2020\|EPI_ISL_424216\|2020-03 | ···················· | ···················· | ···················· | ···················· | ···················· | ···················· |
| A | 590 | hCoV-19/USA/WA-UW-1868/2020\|EPI_ISL_424280\|2020-03 | ···················· | ···················· | ···················· | ···················· | ···················· | ···················· |
| A | 591 | hCoV-19/USA/WA-UW-1864/2020\|EPI_ISL_424281\|2020-03 | ···················· | ···················· | ···················· | ···················· | ···················· | ···················· |
| A | 592 | hCoV-19/USA/WA-UW-1835/2020\|EPI_ISL_424282\|2020-03 | ···················· | ···················· | ···················· | ···················· | ···················· | ···················· |
| A | 593 | hCoV-19/USA/WA-UW-1863/2020\|EPI_ISL_424284\|2020-03 | ···················· | ···················· | ···················· | ···················· | ···················· | ···················· |
| A | 594 | hCoV-19/USA/OR-UW-1851/2020\|EPI_ISL_424285\|2020-03 | ···················· | ···················· | ···················· | ···················· | ···················· | ···················· |
| A | 595 | hCoV-19/USA/OR-UW-1849/2020\|EPI_ISL_424286\|2020-03 | ···················· | ···················· | ···················· | ···················· | ···················· | ···················· |
| A | 596 | hCoV-19/USA/WA-UW-1843/2020\|EPI_ISL_424287\|2020-03 | ···················· | ···················· | ···················· | ···················· | ···················· | ···················· |
| A | 597 | hCoV-19/USA/WA-UW-1846/2020\|EPI_ISL_424288\|2020-03 | ···················· | ···················· | ···················· | ···················· | ···················· | ···················· |
| A | 598 | hCoV-19/USA/WA-UW-1860/2020\|EPI_ISL_424289\|2020-03 | ···················· | ···················· | ···················· | ···················· | ···················· | ···················· |
| A | 599 | hCoV-19/England/NOTT-10E12C/2020\|EPI_ISL_425501\|20 | ···················· | ···················· | ···················· | ···················· | ···················· | ···················· |
| A | 600 | hCoV-19/England/NOTT-10E14A/2020\|EPI_ISL_425502\|20 | ···················· | ···················· | ···················· | ···················· | ···················· | ···················· |
| A | 601 | hCoV-19/England/NOTT-10E186/2020\|EPI_ISL_425504\|20 | ···················· | ···················· | ···················· | ···················· | ···················· | ···················· |
| A | 602 | hCoV-19/England/NOTT-10E195/2020\|EPI_ISL_425505\|20 | ···················· | ···················· | ···················· | ···················· | ···················· | ···················· |
| A | 603 | hCoV-19/England/NOTT-10E1A4/2020\|EPI_ISL_425506\|20 | ···················· | ···················· | ···················· | ···················· | ···················· | ···················· |
| A | 604 | hCoV-19/England/NOTT-10E1B3/2020\|EPI_ISL_425507\|20 | ···················· | ···················· | ···················· | ···················· | ···················· | ···················· |
| A | 605 | hCoV-19/England/NOTT-10E1C2/2020\|EPI_ISL_425508\|20 | ···················· | ···················· | ···················· | ···················· | ···················· | ···················· |
| A | 606 | hCoV-19/England/NOTT-10E1D1/2020\|EPI_ISL_425509\|20 | ···················· | ···················· | ···················· | ···················· | ···················· | ···················· |
| A | 607 | hCoV-19/England/NOTT-10E1E0/2020\|EPI_ISL_425510\|20 | ···················· | ···················· | ···················· | ···················· | ···················· | ···················· |
| A | 608 | hCoV-19/England/NOTT-10E1FF/2020\|EPI_ISL_425511\|20 | ···················· | ···················· | ···················· | ···················· | ···················· | ···················· |
| A | 609 | hCoV-19/England/NOTT-10E20B/2020\|EPI_ISL_425512\|20 | ···················· | ···················· | ···················· | ···················· | ···················· | ···················· |
| A | 610 | hCoV-19/England/NOTT-10E21A/2020\|EPI_ISL_425513\|20 | ···················· | ···················· | ···················· | ···················· | ···················· | ···················· |
| A | 611 | hCoV-19/England/NOTT-10E229/2020\|EPI_ISL_425514\|20 | ···················· | ···················· | ···················· | ···················· | ···················· | ···················· |
| A | 612 | hCoV-19/England/NOTT-10E238/2020\|EPI_ISL_425515\|20 | ···················· | ···················· | ···················· | ···················· | ···················· | ···················· |
| A | 613 | hCoV-19/England/NOTT-10E256/2020\|EPI_ISL_425517\|20 | ···················· | ···················· | ···················· | ···················· | ···················· | ···················· |
| A | 614 | hCoV-19/England/NOTT-10E265/2020\|EPI_ISL_425518\|20 | ···················· | ···················· | ···················· | ···················· | ···················· | ···················· |
| A | 615 | hCoV-19/England/NOTT-10E283/2020\|EPI_ISL_425519\|20 | ···················· | ···················· | ···················· | ···················· | ···················· | ···················· |
| A | 616 | hCoV-19/England/NOTT-10E2B0/2020\|EPI_ISL_425522\|20 | ···················· | ···················· | ···················· | ···················· | ···················· | ···················· |
| A | 617 | hCoV-19/England/NOTT-10E2CF/2020\|EPI_ISL_425523\|20 | ···················· | ···················· | ···················· | ···················· | ···················· | ···················· |
| A | 618 | hCoV-19/England/NOTT-10E2ED/2020\|EPI_ISL_425525\|20 | ···················· | ···················· | ···················· | ···················· | ···················· | ···················· |
| A | 619 | hCoV-19/England/NOTT-10E2FC/2020\|EPI_ISL_425526\|20 | ···················· | ···················· | ···················· | ···················· | ···················· | ···················· |
| A | 620 | hCoV-19/England/NOTT-10E308/2020\|EPI_ISL_425527\|20 | ···················· | ···················· | ···················· | ···················· | ···················· | ···················· |
| A | 621 | hCoV-19/England/NOTT-10E335/2020\|EPI_ISL_425530\|20 | ···················· | ···················· | ···················· | ···················· | ···················· | ···················· |
| A | 622 | hCoV-19/England/NOTT-10E362/2020\|EPI_ISL_425532\|20 | ···················· | ···················· | ···················· | ···················· | ···················· | ···················· |
| A | 623 | hCoV-19/England/NOTT-10E371/2020\|EPI_ISL_425533\|20 | ···················· | ···················· | ···················· | ···················· | ···················· | ···················· |
| A | 624 | hCoV-19/England/NOTT-10E39F/2020\|EPI_ISL_425535\|20 | ···················· | ···················· | ···················· | ···················· | ···················· | ···················· |
| A | 625 | hCoV-19/England/NOTT-10E3AE/2020\|EPI_ISL_425536\|20 | ···················· | ···················· | ···················· | ···················· | ···················· | ···················· |
| A | 626 | hCoV-19/England/NOTT-10E3BD/2020\|EPI_ISL_425537\|20 | ···················· | ···················· | ···················· | ···················· | ···················· | ···················· |
| A | 627 | hCoV-19/England/NOTT-10E423/2020\|EPI_ISL_425544\|20 | ···················· | ···················· | ···················· | ···················· | ···················· | ···················· |
| A | 628 | hCoV-19/England/NOTT-10E450/2020\|EPI_ISL_425547\|20 | ···················· | ···················· | ···················· | ···················· | ···················· | ···················· |
| A | 629 | hCoV-19/Australia/VIC534/2020\|EPI_ISL_426800\|2020- | ···················· | ···················· | ···················· | ···················· | ···················· | ···················· |
| A | 630 | hCoV-19/Australia/VIC535/2020\|EPI_ISL_426801\|2020- | ···················· | ···················· | ···················· | ···················· | ···················· | ···················· |
| A | 631 | hCoV-19/Australia/VIC536/2020\|EPI_ISL_426802\|2020- | ···················· | ···················· | ···················· | ···················· | ···················· | ·R·················· |
| A | 632 | hCoV-19/Australia/VIC537/2020\|EPI_ISL_426803\|2020- | ···················· | ···················· | ···················· | ···················· | ···················· | ···················· |
| A | 633 | hCoV-19/Australia/VIC538/2020\|EPI_ISL_426804\|2020- | ···················· | ···················· | ···················· | ···················· | ···················· | ···················· |
| A | 634 | hCoV-19/Australia/VIC540/2020\|EPI_ISL_426805\|2020- | ···················· | ···················· | ···················· | ···················· | ···················· | ···················· |
| A | 635 | hCoV-19/Australia/VIC541/2020\|EPI_ISL_426806\|2020- | ···················· | ···················· | ···················· | ···················· | ···················· | ···················· |
| A | 636 | hCoV-19/Australia/VIC544/2020\|EPI_ISL_426807\|2020- | ···················· | ···················· | ···················· | ···················· | ···················· | ···················· |
| A | 637 | hCoV-19/Australia/VIC546/2020\|EPI_ISL_426809\|2020- | ···················· | ···················· | ···················· | ···················· | ···················· | ···················· |
| A | 638 | hCoV-19/Australia/VIC547/2020\|EPI_ISL_426810\|2020- | ···················· | ···················· | ···················· | ···················· | ···················· | ···················· |
| A | 639 | hCoV-19/Australia/VIC548/2020\|EPI_ISL_426811\|2020- | ···················· | ···················· | ···················· | ···················· | ···················· | ···················· |
| A | 640 | hCoV-19/Australia/VIC549/2020\|EPI_ISL_426812\|2020- | ···················· | ···················· | ···················· | ···················· | ···················· | ···················· |
| A | 642 | hCoV-19/Australia/VIC553/2020\|EPI_ISL_426814\|2020- | ···················· | ···················· | ···················· | ···················· | ···················· | ···················· |

| | | | | | | | | |
|---|---|---|---|---|---|---|---|---|
| A | 643 | hCoV-19/Australia/VIC555/2020\|EPI_ISL_426816\|2020- | ···················· | ···················· | ···················· | ···················· | ···················· | ···················· |
| A | 645 | hCoV-19/Australia/VIC558/2020\|EPI_ISL_426819\|2020- | ···················· | ···················· | ···················· | ···················· | ···················· | ···················· |
| A | 646 | hCoV-19/Australia/VIC560/2020\|EPI_ISL_426821\|2020- | ···················· | ···················· | ···················· | ···················· | ···················· | ···················· |
| A | 647 | hCoV-19/Australia/VIC561/2020\|EPI_ISL_426822\|2020- | ···················· | ···················· | ···················· | ···················· | ···················· | ···················· |
| A | 649 | hCoV-19/Australia/VIC565/2020\|EPI_ISL_426825\|2020- | ···················· | ···················· | ···················· | ···················· | ···················· | ···················· |
| A | 650 | hCoV-19/Australia/VIC566/2020\|EPI_ISL_426826\|2020- | ···················· | ···················· | ···················· | ···················· | ···················· | ···················· |
| A | 651 | hCoV-19/Australia/VIC567/2020\|EPI_ISL_426827\|2020- | ···················· | ···················· | ···················· | ···················· | ···················· | ···················· |
| A | 652 | hCoV-19/Australia/VIC568/2020\|EPI_ISL_426828\|2020- | ···················· | ···················· | ···················· | ···················· | ···················· | ···················· |
| A | 653 | hCoV-19/Australia/VIC570/2020\|EPI_ISL_426830\|2020- | ···················· | ···················· | ···················· | ···················· | ···················· | ···················· |
| A | 654 | hCoV-19/Australia/VIC571/2020\|EPI_ISL_426831\|2020- | ···················· | ···················· | ···················· | ···················· | ···················· | ···················· |
| A | 655 | hCoV-19/Australia/VIC572/2020\|EPI_ISL_426832\|2020- | ···················· | ···················· | ···················· | ···················· | ···················· | ···················· |
| A | 663 | hCoV-19/Australia/VIC584/2020\|EPI_ISL_426841\|2020- | ···················· | ···················· | ···················· | ···················· | ···················· | ···················· |
| A | 673 | hCoV-19/Australia/VIC597/2020\|EPI_ISL_426854\|2020- | ···················· | ···················· | ···················· | ···················· | ···················· | ···················· |
| A | 674 | hCoV-19/Australia/VIC598/2020\|EPI_ISL_426855\|2020- | ···················· | ···················· | ···················· | ···················· | ···················· | ···················· |
| A | 675 | hCoV-19/Australia/VIC599/2020\|EPI_ISL_426856\|2020- | ···················· | ···················· | ···················· | ···················· | ···················· | ···················· |
| A | 676 | hCoV-19/Australia/VIC603/2020\|EPI_ISL_426858\|2020- | ···················· | ···················· | ···················· | ···················· | ·············Y······ | ···················· |
| A | 677 | hCoV-19/Australia/VIC605/2020\|EPI_ISL_426860\|2020- | ···················· | ···················· | ···················· | ···················· | ···················· | ···················· |
| A | 678 | hCoV-19/Australia/VIC606/2020\|EPI_ISL_426861\|2020- | ···················· | ···················· | ···················· | ···················· | ···················· | ···················· |
| A | 682 | hCoV-19/Australia/VIC612/2020\|EPI_ISL_426866\|2020- | ···················· | ···················· | ···················· | ···················· | ···················· | ···················· |
| A | 684 | hCoV-19/Australia/VIC617/2020\|EPI_ISL_426872\|2020- | ···················· | ···················· | ···················· | ···················· | ···················· | ···················· |
| A | 686 | hCoV-19/Australia/VIC630/2020\|EPI_ISL_426874\|2020- | ···················· | ···················· | ···················· | ···················· | ···················· | ···················· |
| A | 689 | hCoV-19/Australia/VIC636/2020\|EPI_ISL_426877\|2020- | ···················· | ···················· | ···················· | ···················· | ···················· | ···················· |
| A | 691 | hCoV-19/Guangzhou/GZMU0003/2020\|EPI_ISL_429074\|202 | ···················· | ···················· | ···················· | ···················· | ···················· | ···················· |
| A | 692 | hCoV-19/Guangzhou/GZMU0008/2020\|EPI_ISL_429075\|202 | ···················· | ···················· | ···················· | ···················· | ···················· | ···················· |
| A | 693 | hCoV-19/Guangzhou/GZMU0009/2020\|EPI_ISL_429076\|202 | ···················· | ···················· | ···················· | ···················· | ···················· | ···················· |
| A | 694 | hCoV-19/Guangzhou/GZMU0010/2020\|EPI_ISL_429077\|202 | ···················· | ···················· | ···················· | ···················· | ···················· | ···················· |
| A | 695 | hCoV-19/Guangzhou/GZMU0013/2020\|EPI_ISL_429078\|202 | ···················· | ···················· | ···················· | ···················· | ···················· | ···················· |
| A | 696 | hCoV-19/Guangzhou/GZMU0017/2020\|EPI_ISL_429079\|202 | ···················· | ···················· | ···················· | ···················· | ···················· | ···················· |
| A | 697 | hCoV-19/Guangzhou/GZMU0019/2020\|EPI_ISL_429080\|202 | ···················· | ···················· | ···················· | ···················· | ···················· | ···················· |
| A | 698 | hCoV-19/Guangzhou/GZMU0020/2020\|EPI_ISL_429081\|202 | ···················· | ···················· | ···················· | ···················· | ···················· | ···················· |
| A | 699 | hCoV-19/Guangzhou/GZMU0023/2020\|EPI_ISL_429082\|202 | ···················· | ···················· | ···················· | ···················· | ···················· | ···················· |
| A | 700 | hCoV-19/Guangzhou/GZMU0024/2020\|EPI_ISL_429083\|202 | ···················· | ···················· | ···················· | ···················· | ···················· | ···················· |
| A | 701 | hCoV-19/Guangzhou/GZMU0025/2020\|EPI_ISL_429084\|202 | ···················· | ···················· | ···················· | ···················· | ···················· | ···················· |
| A | 702 | hCoV-19/Guangzhou/GZMU0033/2020\|EPI_ISL_429085\|202 | ···················· | ···················· | ···················· | ···················· | ···················· | ···················· |
| A | 703 | hCoV-19/Guangzhou/GZMU0034/2020\|EPI_ISL_429086\|202 | ···················· | ···················· | ···················· | ···················· | ···················· | ···················· |
| A | 704 | hCoV-19/Guangzhou/GZMU0035/2020\|EPI_ISL_429088\|202 | ···················· | ···················· | ···················· | ···················· | ···················· | ···················· |
| A | 705 | hCoV-19/Guangzhou/GZMU0037/2020\|EPI_ISL_429089\|202 | ···················· | ···················· | ···················· | ···················· | ···················· | ···················· |
| A | 706 | hCoV-19/Guangzhou/GZMU0039/2020\|EPI_ISL_429090\|202 | ···················· | ···················· | ···················· | ···················· | ···················· | ···················· |
| A | 707 | hCoV-19/Guangzhou/GZMU0041/2020\|EPI_ISL_429091\|202 | ···················· | ···················· | ···················· | ···················· | ···················· | ···················· |
| A | 708 | hCoV-19/Guangzhou/GZMU0045/2020\|EPI_ISL_429092\|202 | ···················· | ···················· | ···················· | ···················· | ···················· | ···················· |
| A | 709 | hCoV-19/Guangzhou/GZMU0046/2020\|EPI_ISL_429093\|202 | ···················· | ···················· | ···················· | ···················· | ···················· | ···················· |
| A | 710 | hCoV-19/Guangzhou/GZMU0049/2020\|EPI_ISL_429094\|202 | ···················· | ···················· | ···················· | ···················· | ···················· | ···················· |
| A | 711 | hCoV-19/Guangzhou/GZMU0050/2020\|EPI_ISL_429095\|202 | ···················· | ···················· | ···················· | ···················· | ···················· | ···················· |
| A | 712 | hCoV-19/Guangzhou/GZMU0051/2020\|EPI_ISL_429096\|202 | ···················· | ···················· | ···················· | ···················· | ···················· | ···················· |
| A | 713 | hCoV-19/Guangzhou/GZMU0052/2020\|EPI_ISL_429097\|202 | ···················· | ···················· | ···················· | ···················· | ···················· | ···················· |
| A | 714 | hCoV-19/Guangzhou/GZMU0054/2020\|EPI_ISL_429098\|202 | ···················· | ···················· | ···················· | ···················· | ···················· | ···················· |
| A | 715 | hCoV-19/Italy/ABR-IZSGC-TE6571/2020\|EPI_ISL_429231 | ···················· | ···················· | ···················· | ···················· | ···················· | ···················· |
| A | 717 | hCoV-19/DRC/1131/2020\|EPI_ISL_429255\|2020-04-05 | ···················· | ···················· | ···················· | ···················· | ···················· | ···················· |
| A | 718 | hCoV-19/Spain/MD-FISABIO-Vircell001/2020\|EPI_ISL_4 | ···················· | ···················· | ···················· | ···················· | ···················· | ···················· |
| A | 720 | hCoV-19/DRC/1249/2020\|EPI_ISL_429259\|2020-04-06 | ···················· | ···················· | ···················· | ···················· | ···················· | ···················· |
| A | 721 | hCoV-19/Denmark/ALAB-HH01/2020\|EPI_ISL_429262\|2020 | ···················· | ···················· | ···················· | ···················· | ···················· | ···················· |
| A | 722 | hCoV-19/Denmark/ALAB-HH02/2020\|EPI_ISL_429263\|2020 | ···················· | ···················· | ···················· | ···················· | ···················· | ···················· |
| A | 723 | hCoV-19/Denmark/ALAB-HH03/2020\|EPI_ISL_429264\|2020 | ···················· | ···················· | ···················· | ···················· | ···················· | ···················· |
| A | 724 | hCoV-19/Denmark/ALAB-HH04/2020\|EPI_ISL_429265\|2020 | ···················· | ···················· | ···················· | ···················· | ···················· | ···················· |
| A | 725 | hCoV-19/Denmark/ALAB-HH05/2020\|EPI_ISL_429266\|2020 | ···················· | ···················· | ···················· | ···················· | ···················· | ···················· |
| A | 726 | hCoV-19/Denmark/ALAB-HH06/2020\|EPI_ISL_429267\|2020 | ···················· | ···················· | ···················· | ···················· | ···················· | ···················· |
| A | 727 | hCoV-19/Denmark/ALAB-HH07/2020\|EPI_ISL_429268\|2020 | ···················· | ···················· | ···················· | ···················· | ···················· | ···················· |
| A | 728 | hCoV-19/Denmark/ALAB-HH08/2020\|EPI_ISL_429269\|2020 | ···················· | ···················· | ···················· | ···················· | ···················· | ···················· |
| A | 729 | hCoV-19/USA/UT-0511/2020\|EPI_ISL_430058\|2020-04-03 | ···················· | ···················· | ···················· | ···················· | ···················· | ···················· |
| A | 730 | hCoV-19/USA/UT-0513/2020\|EPI_ISL_430059\|2020-04-03 | ···················· | ···················· | ···················· | ···················· | ···················· | ···················· |
| A | 732 | hCoV-19/USA/UT-0519/2020\|EPI_ISL_430061\|2020-04-03 | ···················· | ···················· | ···················· | ···················· | ···················· | ···················· |
| A | 733 | hCoV-19/USA/UT-0520/2020\|EPI_ISL_430062\|2020-04-03 | ···················· | ···················· | ···················· | ···················· | ···················· | ···················· |
| A | 734 | hCoV-19/Australia/VIC-CBA4/2020\|EPI_ISL_430064\|202 | ···················· | ···················· | ···················· | ···················· | ···················· | ···················· |
| A | 735 | hCoV-19/Australia/VIC-CBA5/2020\|EPI_ISL_430065\|202 | ···················· | ···················· | ···················· | ···················· | ···················· | ···················· |
| A | 736 | hCoV-19/Russia/StPetersburg-RII4705S/2020\|EPI_ISL_ | ···················· | ···················· | ···················· | ···················· | ···················· | ···················· |
| A | 737 | hCoV-19/Russia/StPetersburg-RII4712V/2020\|EPI_ISL_ | ···················· | ···················· | ···················· | ···················· | ···················· | ···················· |
| A | 738 | hCoV-19/Russia/StPetersburg-RII4780S/2020\|EPI_ISL_ | ···················· | ···················· | ···················· | ···················· | ···················· | ···················· |
| A | 739 | hCoV-19/Russia/StPetersburg-RII4917S/2020\|EPI_ISL_ | ···················· | ···················· | ···················· | ···················· | ···················· | ···················· |
| A | 740 | hCoV-19/Russia/StPetersburg-RII4936S/2020\|EPI_ISL_ | ···················· | ···················· | ···················· | ···················· | ···················· | ···················· |
| A | 741 | hCoV-19/Russia/StPetersburg-RII4937S/2020\|EPI_ISL_ | ···················· | ···················· | ···················· | ···················· | ···················· | ···················· |
| A | 742 | hCoV-19/Russia/StPetersburg-RII4938S/2020\|EPI_ISL_ | ···················· | ···················· | ···················· | ···················· | ···················· | ···················· |
| A | 743 | hCoV-19/Russia/StPetersburg-RII4944S/2020\|EPI_ISL_ | ···················· | ···················· | ···················· | ···················· | ···················· | ···················· |
| A | 744 | hCoV-19/Russia/StPetersburg-RII4949S/2020\|EPI_ISL_ | ···················· | ···················· | ···················· | ···················· | ···················· | ···················· |
| A | 745 | hCoV-19/Russia/StPetersburg-RII4969S/2020\|EPI_ISL_ | ···················· | ···················· | ···················· | ···················· | ···················· | ···················· |
| A | 746 | hCoV-19/Russia/StPetersburg-RII4970S/2020\|EPI_ISL_ | ···················· | ···················· | ···················· | ···················· | ···················· | ···················· |
| A | 747 | hCoV-19/Russia/StPetersburg-RII4972S/2020\|EPI_ISL_ | ···················· | ···················· | ···················· | ···················· | ···················· | ···················· |
| A | 748 | hCoV-19/Russia/StPetersburg-RII4980S/2020\|EPI_ISL_ | ···················· | ···················· | ···················· | ···················· | ···················· | ···················· |
| A | 749 | hCoV-19/Russia/StPetersburg-RII4983S/2020\|EPI_ISL_ | ···················· | ···················· | ···················· | ···················· | ···················· | ···················· |

```
A 750 hCoV-19/Russia/StPetersburg-RII4984S/2020|EPI_ISL_ .................... .................... .................... .................... .................... ....................
A 751 hCoV-19/Russia/StPetersburg-RII4985S/2020|EPI_ISL_ .................... .................... .................... .................... .................... ....................
A 752 hCoV-19/Russia/StPetersburg-RII4988S/2020|EPI_ISL_ .................... .................... .................... .................... .................... ....................
A 753 hCoV-19/Russia/StPetersburg-RII5017V/2020|EPI_ISL_ .................... .................... .................... .................... .................... ....................
A 754 hCoV-19/Russia/StPetersburg-RII5029S/2020|EPI_ISL_ .................... .................... .................... .................... .................... ....................
A 755 hCoV-19/Russia/StPetersburg-RII5033S/2020|EPI_ISL_ .................... .................... .................... .................... .................... ....................
A 756 hCoV-19/Russia/StPetersburg-RII5047S/2020|EPI_ISL_ .................... .................... .................... .................... .................... ....................
A 757 hCoV-19/Russia/StPetersburg-RII5169S/2020|EPI_ISL_ .................... .................... .................... .................... .................... ....................
A 758 hCoV-19/Russia/StPetersburg-RII5170S/2020|EPI_ISL_ .................... .................... .................... .................... .................... ....................
A 759 hCoV-19/Russia/StPetersburg-RII5244S/2020|EPI_ISL_ .................... .................... .................... .................... .................... ....................
A 760 hCoV-19/Russia/StPetersburg-RII5247S/2020|EPI_ISL_ .................... .................... .................... .................... .................... ....................
A 761 hCoV-19/Russia/StPetersburg-RII5633S/2020|EPI_ISL_ .................... .................... .................... .................... .................... ....................
A 762 hCoV-19/Russia/StPetersburg-RII5634S/2020|EPI_ISL_ .................... .................... .................... .................... .................... ....................
A 763 hCoV-19/Russia/StPetersburg-RII5635S/2020|EPI_ISL_ .................... .................... .................... .................... .................... ....................
A 764 hCoV-19/Russia/StPetersburg-RII5642S/2020|EPI_ISL_ .................... .................... .................... .................... .................... ....................
A 765 hCoV-19/Russia/StPetersburg-RII5643S/2020|EPI_ISL_ .................... .................... .................... .................... .................... ....................
A 766 hCoV-19/Russia/StPetersburg-RII5644S/2020|EPI_ISL_ .................... .................... .................... .................... .................... ....................
A 767 hCoV-19/Russia/StPetersburg-RII5654S/2020|EPI_ISL_ .................... .................... .................... .................... .................... ....................
A 768 hCoV-19/Russia/StPetersburg-RII5942S/2020|EPI_ISL_ .................... .................... .................... .................... .................... ....................
A 769 hCoV-19/Guangdong/20SF174/2020|EPI_ISL_406531|2020 .................... .................... .................... .................... .................... ....................
A 770 hCoV-19/Guangzhou/20SF206/2020|EPI_ISL_406533|2020 .................... .................... .................... .................... .................... ....................
A 771 hCoV-19/Foshan/20SF207/2020|EPI_ISL_406534|2020-01 .................... .A.................. .................... .................... .................... ....................
A 772 hCoV-19/Foshan/20SF210/2020|EPI_ISL_406535|2020-01 .................... .................... .................... .................... .................... ....................
A 773 hCoV-19/Foshan/20SF211/2020|EPI_ISL_406536|2020-01 .................... .................... .................... .................... .................... ....................
A 774 hCoV-19/Guangdong/20SF201/2020|EPI_ISL_406538|2020 .................... .................... .................... .................... .................... ....................
A 775 hCoV-19/Shenzhen/SZTH-001/2020|EPI_ISL_406592|2020 .................... .................... .................... .................... .................... ....................
A 776 hCoV-19/Shenzhen/SZTH-002/2020|EPI_ISL_406593|2020 .................... .................... .................... .................... .................... ....................
A 777 hCoV-19/France/IDF-0372/2020|EPI_ISL_406596|2020-0 .................... .................... .................... .................... .................... ....................
A 778 hCoV-19/France/IDF-0373/2020|EPI_ISL_406597|2020-0 .................... .................... .................... .................... .................... ....................
A 779 hCoV-19/Australia/NSW01/2020|EPI_ISL_407893|2020-0 .................... .................... .................... .................... .................... ....................
A 780 hCoV-19/Australia/QLD01/2020|EPI_ISL_407894|2020-0 .................... .................... .................... .................... .................... ....................
A 781 hCoV-19/Australia/QLD02/2020|EPI_ISL_407896|2020-0 .................... .................... .................... .................... .................... ....................
A 782 hCoV-19/Weifang/WF0014/2020|EPI_ISL_413711|2020-02 .................... .................... .................... .................... .................... ....................
A 783 hCoV-19/USA/WI-UW-06/2020|EPI_ISL_417200|2020-03-2 .................... .................... .................... .................... .................... ....................
A 784 hCoV-19/USA/WI-UW-07/2020|EPI_ISL_417201|2020-03-2 .................... .................... .................... .................... .................... ....................
A 785 hCoV-19/USA/WI-UW-08/2020|EPI_ISL_417202|2020-03-2 .................... .................... .................... .................... .................... ....................
A 786 hCoV-19/USA/WI-UW-09/2020|EPI_ISL_417203|2020-03-2 .................... .................... .................... .................... .................... ....................
A 787 hCoV-19/USA/WI-UW-10/2020|EPI_ISL_417204|2020-03-2 .................... .................... .................... .................... .................... ....................
A 788 hCoV-19/New Zealand/CoV001/2020|EPI_ISL_417211|202 .................... .................... .................... .................... .................... ....................
A 789 hCoV-19/New Zealand/CoV002/2020|EPI_ISL_417212|202 .................... .................... .................... .................... .................... ....................
A 790 hCoV-19/England/20102068502/2020|EPI_ISL_417213|20 .................... .................... .................... .................... .................... ....................
A 793 hCoV-19/England/20104002606/2020|EPI_ISL_417220|20 .................... .................... .................... .................... .................... ....................
A 794 hCoV-19/England/20104003002/2020|EPI_ISL_417222|20 .................... .................... .................... .................... .................... ....................
A 795 hCoV-19/England/20104004902/2020|EPI_ISL_417226|20 .................... .................... .................... .................... .................... ....................
A 796 hCoV-19/England/20104007503/2020|EPI_ISL_417227|20 .................... .................... .................... .................... .................... ....................
A 797 hCoV-19/England/20104008402/2020|EPI_ISL_417228|20 .................... .................... .................... .................... .................... ....................
A 798 hCoV-19/England/20104008702/2020|EPI_ISL_417230|20 .................... .................... .................... .................... .................... ....................
A 799 hCoV-19/England/20104008802/2020|EPI_ISL_417231|20 .................... .................... .................... .................... .................... ....................
A 800 hCoV-19/England/20104008902/2020|EPI_ISL_417232|20 .................... .................... .................... .................... .................... ....................
A 801 hCoV-19/England/20104009002/2020|EPI_ISL_417233|20 .................... .................... .................... .................... .................... ....................
A 802 hCoV-19/England/20104009102/2020|EPI_ISL_417234|20 .................... .................... .................... .................... .................... ....................
A 803 hCoV-19/England/20104013703/2020|EPI_ISL_417235|20 .................... .................... .................... .................... .................... ....................
A 804 hCoV-19/England/20104015302/2020|EPI_ISL_417236|20 .................... .................... .................... .................... .................... ....................
A 805 hCoV-19/England/20104035803/2020|EPI_ISL_417238|20 .................... ......C............. .................... .................... .................... ....................
A 806 hCoV-19/England/20106003303/2020|EPI_ISL_417239|20 .................... .................... .................... .................... .................... ....................
A 807 hCoV-19/England/20106004803/2020|EPI_ISL_417240|20 .................... .................... .................... .................... .................... ....................
A 808 hCoV-19/England/20106005403/2020|EPI_ISL_417244|20 .................... .................... .................... .................... .................... ....................
A 809 hCoV-19/England/20106145903/2020|EPI_ISL_417246|20 .................... .................... .................... .................... .................... ....................
A 810 hCoV-19/England/20108003302/2020|EPI_ISL_417248|20 .................... .................... .................... .................... .................... ....................
A 811 hCoV-19/England/20109051806/2020|EPI_ISL_417270|20 .................... .................... .................... .................... .................... ....................
A 812 hCoV-19/England/20109052006/2020|EPI_ISL_417272|20 .................... .................... .................... .................... .................... ....................
A 813 hCoV-19/England/20109052106/2020|EPI_ISL_417273|20 .................... .................... .................... .................... .................... ....................
A 814 hCoV-19/England/20109052506/2020|EPI_ISL_417276|20 .................... .................... .................... .................... .................... ....................
A 815 hCoV-19/England/20109053406/2020|EPI_ISL_417278|20 .................... .................... .................... .................... .................... ....................
A 816 hCoV-19/England/20109053606/2020|EPI_ISL_417279|20 .................... .................... .................... .................... .................... ....................
A 817 hCoV-19/England/20109054806/2020|EPI_ISL_417280|20 .................... .................... .................... .................... .................... ....................
A 818 hCoV-19/England/20109094106/2020|EPI_ISL_417290|20 .................... .................... .................... .................... .................... ....................
A 819 hCoV-19/England/20109094206/2020|EPI_ISL_417291|20 .................... .................... .................... .................... .................... ....................
A 820 hCoV-19/England/20109094506/2020|EPI_ISL_417292|20 .................... .................... .................... .................... .................... ....................
A 821 hCoV-19/England/20109097506/2020|EPI_ISL_417293|20 .................... .................... .................... .................... .................... ....................
A 822 hCoV-19/England/20109098806/2020|EPI_ISL_417295|20 .................... .................... .................... .................... .................... ....................
A 823 hCoV-19/England/20109098906/2020|EPI_ISL_417296|20 .................... .................... .................... .................... .................... ....................
A 824 hCoV-19/England/20109099006/2020|EPI_ISL_417297|20 .................... .................... .................... .................... .................... ....................
A 825 hCoV-19/England/20109099106/2020|EPI_ISL_417298|20 .................... .................... .................... .................... .................... ....................
A 826 hCoV-19/England/20109099206/2020|EPI_ISL_417299|20 .................... .................... .................... .................... .................... ....................
A 827 hCoV-19/Hangzhou/HZ60/2020|EPI_ISL_418502|2020-01- .................... .................... .................... .................... .................... ....................
A 828 hCoV-19/Hangzhou/HZ62/2020|EPI_ISL_418503|2020-01- .................... .................... .................... .................... .................... ....................
A 829 hCoV-19/Hangzhou/HZ79/2020|EPI_ISL_418504|2020-01- .................... .................... .................... .................... .................... ....................
```

| | | | | | | | |
|---|---|---|---|---|---|---|---|
| A | 830 hCoV-19/Hangzhou/HZ90/2020\|EPI_ISL_418506\|2020-01- | ···················· | ···················· | ····················· | ···················· | ···················· | ···················· |
| A | 831 hCoV-19/Hangzhou/HZ91/2020\|EPI_ISL_418507\|2020-01- | ···················· | ···················· | ····················· | ···················· | ···················· | ···················· |
| A | 832 hCoV-19/Hangzhou/HZ162/2020\|EPI_ISL_418508\|2020-01 | ···················· | ···················· | ····················· | ···················· | ···················· | ···················· |
| A | 833 hCoV-19/Hangzhou/HZ178/2020\|EPI_ISL_418509\|2020-01 | ···················· | ···················· | ····················· | ···················· | ···················· | ···················· |
| A | 834 hCoV-19/Hangzhou/HZ185/2020\|EPI_ISL_418510\|2020-01 | ···················· | ···················· | ····················· | ···················· | ···················· | ···················· |
| A | 835 hCoV-19/Hangzhou/HZ477/2020\|EPI_ISL_418511\|2020-01 | ···················· | ···················· | ····················· | ···················· | ···················· | ···················· |
| A | 836 hCoV-19/Hangzhou/HZ481/2020\|EPI_ISL_418512\|2020-01 | ···················· | ···················· | ····················· | ···················· | ···················· | ···················· |
| A | 837 hCoV-19/Hangzhou/HZ551/2020\|EPI_ISL_418513\|2020-01 | ···················· | ···················· | ····················· | ···················· | ···················· | ···················· |
| A | 838 hCoV-19/Hangzhou/HZ576/2020\|EPI_ISL_418514\|2020-01 | ···················· | ···················· | ····················· | ···················· | ···················· | ···················· |
| A | 839 hCoV-19/Hangzhou/HZ638/2020\|EPI_ISL_418515\|2020-01 | ···················· | ···················· | ····················· | ···················· | ···················· | ···················· |
| A | 840 hCoV-19/Ireland/TA-NVRL-21023/2020\|EPI_ISL_418516\| | ···················· | ···················· | ····················· | ···················· | ···················· | ···················· |
| A | 841 hCoV-19/Ireland/D-NVRL-21145/2020\|EPI_ISL_418548\|2 | ···················· | ···················· | ····················· | ···················· | ···················· | ···················· |
| A | 842 hCoV-19/Australia/VIC135/2020\|EPI_ISL_419828\|2020- | ···················· | ···················· | ····················· | ···················· | ···················· | ···················· |
| A | 843 hCoV-19/Australia/VIC136/2020\|EPI_ISL_419829\|2020- | ···················· | ···················· | ····················· | ···················· | ···················· | ···················· |
| A | 844 hCoV-19/Australia/VIC137/2020\|EPI_ISL_419830\|2020- | ···················· | ···················· | ····················· | ···················· | ···················· | ···················· |
| A | 845 hCoV-19/Australia/NT01/2020\|EPI_ISL_419831\|2020 | ···················· | ···················· | ····················· | ···················· | ···················· | ···················· |
| A | 846 hCoV-19/Australia/NT02/2020\|EPI_ISL_419832\|2020 | ···················· | ···················· | ····················· | ···················· | ···················· | ···················· |
| A | 847 hCoV-19/Australia/VIC138/2020\|EPI_ISL_419834\|2020- | ···················· | ···················· | ····················· | ···················· | ···················· | ···················· |
| A | 848 hCoV-19/Australia/NT05/2020\|EPI_ISL_419835\|2020 | ···················· | ···················· | ····················· | ···················· | ···················· | ···················· |
| A | 849 hCoV-19/Australia/VIC139/2020\|EPI_ISL_419836\|2020- | ···················· | ···················· | ····················· | ···················· | ···················· | ···················· |
| A | 850 hCoV-19/Australia/VIC140/2020\|EPI_ISL_419837\|2020- | ···················· | ···················· | ····················· | ···················· | ···················· | ···················· |
| A | 851 hCoV-19/Australia/VIC141/2020\|EPI_ISL_419838\|2020- | ···················· | ···················· | ····················· | ···················· | ···················· | ···················· |
| A | 852 hCoV-19/Australia/VIC142/2020\|EPI_ISL_419839\|2020- | ···················· | ···················· | ····················· | ···················· | ···················· | ···················· |
| A | 853 hCoV-19/Australia/VIC143/2020\|EPI_ISL_419840\|2020- | ···················· | ···················· | ····················· | ···················· | ···················· | ···················· |
| A | 854 hCoV-19/Australia/VIC144/2020\|EPI_ISL_419841\|2020- | ···················· | ···················· | ····················· | ···················· | ···················· | ···················· |
| A | 855 hCoV-19/Australia/VIC145/2020\|EPI_ISL_419842\|2020- | ···················· | ···················· | ····················· | ···················· | ···················· | ···················· |
| A | 856 hCoV-19/Australia/VIC146/2020\|EPI_ISL_419843\|2020- | ···················· | ···················· | ····················· | ···················· | ···················· | ···················· |
| A | 857 hCoV-19/Australia/VIC147/2020\|EPI_ISL_419844\|2020- | ···················· | ···················· | ····················· | ···················· | ···················· | ···················· |
| A | 858 hCoV-19/Australia/VIC148/2020\|EPI_ISL_419845\|2020- | ···················· | ···················· | ····················· | ···················· | ···················· | ···················· |
| A | 859 hCoV-19/Australia/VIC149/2020\|EPI_ISL_419846\|2020- | ···················· | ···················· | ····················· | ···················· | ···················· | ···················· |
| A | 860 hCoV-19/Australia/VIC150/2020\|EPI_ISL_419847\|2020- | ···················· | ···················· | ····················· | ···················· | ···················· | ···················· |
| A | 861 hCoV-19/Australia/VIC151/2020\|EPI_ISL_419848\|2020- | ···················· | ···················· | ····················· | ···················· | ···················· | ···················· |
| A | 862 hCoV-19/Australia/VIC152/2020\|EPI_ISL_419849\|2020- | ···················· | ···················· | ····················· | ···················· | ···················· | ···················· |
| A | 863 hCoV-19/Australia/VIC153/2020\|EPI_ISL_419850\|2020- | ···················· | ···················· | ····················· | ···················· | ···················· | ···················· |
| A | 864 hCoV-19/Australia/VIC154/2020\|EPI_ISL_419851\|2020- | ···················· | ···················· | ····················· | ···················· | ···················· | ···················· |
| A | 865 hCoV-19/Australia/VIC155/2020\|EPI_ISL_419852\|2020- | ···················· | ···················· | ····················· | ···················· | ···················· | ···················· |
| A | 866 hCoV-19/Australia/VIC156/2020\|EPI_ISL_419853\|2020- | ···················· | ···················· | ····················· | ···················· | ···················· | ···················· |
| A | 867 hCoV-19/Australia/VIC157/2020\|EPI_ISL_419854\|2020- | ···················· | ···················· | ····················· | ···················· | ···················· | ···················· |
| A | 868 hCoV-19/Australia/VIC158/2020\|EPI_ISL_419855\|2020- | ···················· | ···················· | ····················· | ···················· | ···················· | ···················· |
| A | 869 hCoV-19/Australia/VIC159/2020\|EPI_ISL_419856\|2020- | ···················· | ···················· | ····················· | ···················· | ···················· | ···················· |
| A | 870 hCoV-19/Australia/VIC160/2020\|EPI_ISL_419857\|2020- | ···················· | ···················· | ····················· | ···················· | ···················· | ···················· |
| A | 871 hCoV-19/Australia/VIC162/2020\|EPI_ISL_419859\|2020- | ···················· | ···················· | ····················· | ···················· | ···················· | ···················· |
| A | 872 hCoV-19/Australia/VIC163/2020\|EPI_ISL_419860\|2020- | ···················· | ···················· | ····················· | ···················· | ···················· | ···················· |
| A | 873 hCoV-19/Australia/VIC164/2020\|EPI_ISL_419861\|2020- | ···················· | ···················· | ····················· | ···················· | ···················· | ···················· |
| A | 874 hCoV-19/Australia/VIC165/2020\|EPI_ISL_419862\|2020- | ···················· | ···················· | ····················· | ···················· | ···················· | ···················· |
| A | 875 hCoV-19/Australia/VIC166/2020\|EPI_ISL_419863\|2020- | ···················· | ···················· | ····················· | ···················· | ···················· | ···················· |
| A | 876 hCoV-19/Australia/VIC167/2020\|EPI_ISL_419864\|2020- | ···················· | ···················· | ····················· | ···················· | ···················· | ···················· |
| A | 877 hCoV-19/Australia/VIC168/2020\|EPI_ISL_419865\|2020- | ···················· | ···················· | ····················· | ···················· | ···················· | ···················· |
| A | 878 hCoV-19/Australia/VIC169/2020\|EPI_ISL_419866\|2020- | ···················· | ·············A······ | ····················· | ···················· | ···················· | ···················· |
| A | 879 hCoV-19/Australia/VIC170/2020\|EPI_ISL_419867\|2020- | ···················· | ···················· | ····················· | ···················· | ···················· | ···················· |
| A | 880 hCoV-19/Australia/VIC171/2020\|EPI_ISL_419868\|2020- | ···················· | ···················· | ····················· | ···················· | ···················· | ···················· |
| A | 881 hCoV-19/Australia/VIC172/2020\|EPI_ISL_419869\|2020- | ···················· | ···················· | ····················· | ···················· | ···················· | ···················· |
| A | 882 hCoV-19/Australia/VIC173/2020\|EPI_ISL_419870\|2020- | ···················· | ···················· | ····················· | ···················· | ···················· | ···················· |
| A | 883 hCoV-19/Australia/VIC174/2020\|EPI_ISL_419871\|2020- | ···················· | ···················· | ····················· | ···················· | ···················· | ···················· |
| A | 884 hCoV-19/Australia/VIC175/2020\|EPI_ISL_419872\|2020- | ···················· | ···················· | ····················· | ···················· | ···················· | ···················· |
| A | 885 hCoV-19/Australia/VIC176/2020\|EPI_ISL_419873\|2020- | ···················· | ···················· | ····················· | ···················· | ···················· | ···················· |
| A | 886 hCoV-19/Australia/VIC177/2020\|EPI_ISL_419874\|2020- | ···················· | ···················· | ····················· | ···················· | ···················· | ···················· |
| A | 887 hCoV-19/Australia/VIC178/2020\|EPI_ISL_419875\|2020- | ···················· | ···················· | ····················· | ···················· | ···················· | ···················· |
| A | 888 hCoV-19/Australia/VIC179/2020\|EPI_ISL_419876\|2020- | ···················· | ···················· | ····················· | ···················· | ···················· | ···················· |
| A | 889 hCoV-19/Australia/VIC180/2020\|EPI_ISL_419877\|2020- | ···················· | ···················· | ····················· | ···················· | ···················· | ···················· |
| A | 890 hCoV-19/Australia/VIC181/2020\|EPI_ISL_419878\|2020- | ···················· | ···················· | ····················· | ···················· | ···················· | ···················· |
| A | 891 hCoV-19/Australia/VIC183/2020\|EPI_ISL_419879\|2020- | ···················· | ···················· | ····················· | ···················· | ···················· | ···················· |
| A | 892 hCoV-19/Australia/VIC184/2020\|EPI_ISL_419880\|2020- | ···················· | ···················· | ····················· | ···················· | ···················· | ···················· |
| A | 893 hCoV-19/Australia/VIC185/2020\|EPI_ISL_419881\|2020- | ···················· | ···················· | ····················· | ···················· | ···················· | ···················· |
| A | 894 hCoV-19/Australia/VIC186/2020\|EPI_ISL_419882\|2020- | ···················· | ···················· | ····················· | ···················· | ···················· | ···················· |
| A | 895 hCoV-19/Australia/VIC187/2020\|EPI_ISL_419883\|2020- | ···················· | ···················· | ····················· | ···················· | ···················· | ···················· |
| A | 896 hCoV-19/Australia/VIC188/2020\|EPI_ISL_419884\|2020- | ···················· | ···················· | ····················· | ···················· | ···················· | ···················· |
| A | 897 hCoV-19/Australia/VIC189/2020\|EPI_ISL_419885\|2020- | ···················· | ···················· | ····················· | ···················· | ···················· | ···················· |
| A | 898 hCoV-19/Australia/VIC190/2020\|EPI_ISL_419886\|2020- | ···················· | ···················· | ····················· | ···················· | ···················· | ···················· |
| A | 899 hCoV-19/Australia/VIC191/2020\|EPI_ISL_419887\|2020- | ···················· | ···················· | ····················· | ···················· | ···················· | ···················· |
| A | 900 hCoV-19/Australia/VIC192/2020\|EPI_ISL_419888\|2020- | ···················· | ···················· | ····················· | ···················· | ···················· | ···················· |
| A | 901 hCoV-19/Australia/VIC193/2020\|EPI_ISL_419889\|2020- | ···················· | ···················· | ····················· | ···················· | ···················· | ···················· |
| A | 902 hCoV-19/Australia/VIC194/2020\|EPI_ISL_419890\|2020- | ·······C············ | ···················· | ····················· | ···················· | ···················· | ···················· |
| A | 903 hCoV-19/Australia/VIC195/2020\|EPI_ISL_419891\|2020- | ···················· | ···················· | ····················· | ···················· | ···················· | ···················· |
| A | 904 hCoV-19/Australia/VIC196/2020\|EPI_ISL_419892\|2020- | ···················· | ···················· | ····················· | ···················· | ···················· | ···················· |
| A | 905 hCoV-19/Australia/VIC197/2020\|EPI_ISL_419893\|2020- | ···················· | ···················· | ····················· | ···················· | ···················· | ···················· |
| A | 906 hCoV-19/Australia/VIC198/2020\|EPI_ISL_419894\|2020- | ···················· | ···················· | ····················· | ···················· | ···················· | ···················· |
| A | 907 hCoV-19/Australia/VIC199/2020\|EPI_ISL_419895\|2020- | ···················· | ···················· | ····················· | ···················· | ···················· | ···················· |

| | | | | | | | | |
|---|---|---|---|---|---|---|---|---|
| A | 908 hCoV-19/Australia/VIC201/2020\|EPI_ISL_419896\|2020- | .................... | .................... | .................... | .................... | .................... | .................... | |
| A | 909 hCoV-19/Australia/VIC202/2020\|EPI_ISL_419897\|2020- | .................... | .................... | .................... | .................... | .................... | .................... | |
| A | 910 hCoV-19/Australia/VIC203/2020\|EPI_ISL_419898\|2020- | .................... | .................... | .................... | .................... | .................... | .................... | |
| A | 911 hCoV-19/Australia/VIC204/2020\|EPI_ISL_419899\|2020- | .................... | .................... | .................... | .................... | .................... | .................... | |
| A | 912 hCoV-19/USA/UT-0290/2020\|EPI_ISL_420810\|2020-03-27 | .................... | .................... | .................... | .................... | .................... | .................... | |
| A | 913 hCoV-19/USA/UT-0297/2020\|EPI_ISL_420812\|2020-03-27 | .................... | .................... | .................... | .................... | .................... | .................... | |
| A | 914 hCoV-19/USA/UT-0301/2020\|EPI_ISL_420814\|2020-03-27 | .................... | .................... | .................... | .................... | .................... | .................... | |
| A | 915 hCoV-19/USA/UT-0347/2020\|EPI_ISL_420820\|2020-03-25 | .................... | .................... | .................... | .................... | .................... | .................... | |
| A | 916 hCoV-19/USA/UT-0348/2020\|EPI_ISL_420821\|2020-03-26 | .................... | .................... | .................... | .................... | .................... | .................... | |
| A | 917 hCoV-19/USA/UT-0349/2020\|EPI_ISL_420822\|2020-03-25 | .................... | .................... | .................... | .................... | .................... | .................... | |
| A | 918 hCoV-19/USA/UT-0352/2020\|EPI_ISL_420824\|2020-03-26 | .................... | .................... | .................... | .................... | .................... | .................... | |
| A | 919 hCoV-19/USA/UT-0361/2020\|EPI_ISL_420825\|2020-03-25 | .................... | .................... | .................... | .................... | .................... | .................... | |
| A | 920 hCoV-19/DRC/253/2020\|EPI_ISL_420838\|2020-03-22 | .................... | .................... | .................... | .................... | .................... | .................... | |
| A | 921 hCoV-19/DRC/254/2020\|EPI_ISL_420839\|2020-03-22 | .................... | .................... | .................... | .................... | .................... | .................... | |
| A | 922 hCoV-19/DRC/299/2020\|EPI_ISL_420840\|2020-03-20 | .................... | .................... | .................... | .................... | .................... | .................... | |
| A | 923 hCoV-19/DRC/300/2020\|EPI_ISL_420841\|2020-03-22 | .................... | .................... | .................... | .................... | .................... | .................... | |
| A | 924 hCoV-19/DRC/307/2020\|EPI_ISL_420842\|2020-03 | .................... | .................... | .................... | .................... | .................... | .................... | |
| A | 925 hCoV-19/DRC/353/2020\|EPI_ISL_420843\|2020-03 | .................... | .................... | .................... | .................... | .................... | .................... | |
| A | 926 hCoV-19/DRC/355/2020\|EPI_ISL_420844\|2020-03 | .................... | .................... | .................... | .................... | .................... | .................... | |
| A | 927 hCoV-19/DRC/397/2020\|EPI_ISL_420845\|2020-03-26 | .................... | .................... | .................... | .................... | .................... | .................... | |
| A | 928 hCoV-19/DRC/400/2020\|EPI_ISL_420846\|2020-03 | .................... | .................... | .................... | .................... | .................... | .................... | |
| A | 929 hCoV-19/DRC/431/2020\|EPI_ISL_420847\|2020-03-26 | .................... | .................... | .................... | .................... | .................... | .................... | |
| A | 930 hCoV-19/DRC/445/2020\|EPI_ISL_420848\|2020-03 | .................... | .................... | .................... | .................... | .................... | .................... | |
| A | 931 hCoV-19/DRC/523/2020\|EPI_ISL_420849\|2020-03-28 | .................... | .................... | .................... | .................... | .................... | .................... | |
| A | 932 hCoV-19/DRC/352/2020\|EPI_ISL_420850\|2020-03 | .................... | .................... | .................... | .................... | .................... | .................... | |
| A | 933 hCoV-19/DRC/376/2020\|EPI_ISL_420851\|2020-03-25 | .................... | .................... | .................... | .................... | .................... | .................... | |
| A | 934 hCoV-19/DRC/396/2020\|EPI_ISL_420852\|2020-03-26 | .................... | .................... | .................... | .................... | .................... | .................... | |
| A | 935 hCoV-19/DRC/402/2020\|EPI_ISL_420853\|2020-03-26 | .................... | .................... | .................... | .................... | .................... | .................... | |
| A | 936 hCoV-19/DRC/521/2020\|EPI_ISL_420854\|2020-03-25 | .................... | .................... | .................... | .................... | .................... | .................... | |
| A | 937 hCoV-19/Australia/VIC-CBA1/2020\|EPI_ISL_420855\|202 | .................... | .................... | .................... | .................... | .................... | .................... | |
| A | 938 hCoV-19/Australia/VIC-CBA2/2020\|EPI_ISL_420876\|202 | .................... | .................... | .................... | .................... | .................... | .................... | |
| A | 939 hCoV-19/Australia/VIC-CBA3/2020\|EPI_ISL_420877\|202 | .................... | .................... | .................... | .................... | .................... | .................... | |
| A | 940 hCoV-19/Australia/QLDID929/2020\|EPI_ISL_420878\|202 | .................... | .................... | .................... | .................... | .................... | .................... | |
| A | 941 hCoV-19/Australia/QLDID931/2020\|EPI_ISL_420879\|202 | .................... | .................... | .................... | .................... | .................... | .................... | |
| A | 942 hCoV-19/Japan/Hu_DP_Kng_19-031/2020\|EPI_ISL_420889 | .................... | .................... | .................... | .................... | .................... | .................... | |
| A | 943 hCoV-19/Japan/Hu_Kng_19-437/2020\|EPI_ISL_420890\|20 | .................... | .................... | .................... | .................... | .................... | .................... | |
| A | 944 hCoV-19/USA/WA-UW-1569/2020\|EPI_ISL_423021\|2020-03 | .................... | .................... | .................... | .................... | .................... | .................... | |
| A | 945 hCoV-19/USA/WA-UW-1562/2020\|EPI_ISL_423022\|2020-03 | .................... | .................... | .................... | .................... | .................... | .................... | |
| A | 946 hCoV-19/USA/UN-UW-1449/2020\|EPI_ISL_423023\|2020-03 | .................... | .................... | .................... | .................... | .................... | .................... | |
| A | 947 hCoV-19/USA/WA-UW-1435/2020\|EPI_ISL_423024\|2020-03 | .................... | .................... | .................... | .................... | .................... | .................... | |
| A | 948 hCoV-19/USA/WA-UW-1441/2020\|EPI_ISL_423025\|2020-03 | .................... | .................... | .................... | .................... | .................... | .................... | |
| A | 949 hCoV-19/USA/WA-UW-1440/2020\|EPI_ISL_423026\|2020-03 | .................... | .................... | .................... | .................... | .................... | .................... | |
| A | 950 hCoV-19/USA/WA-UW-1470/2020\|EPI_ISL_423028\|2020-03 | .................... | .................... | .................... | .................... | .................... | .................... | |
| A | 951 hCoV-19/England/20134016802/2020\|EPI_ISL_423091\|20 | .................... | .................... | .................... | .................... | .................... | .................... | |
| A | 952 hCoV-19/England/20134016902/2020\|EPI_ISL_423092\|20 | .................... | .................... | .................... | .................... | .................... | .................... | |
| A | 954 hCoV-19/England/20134017504/2020\|EPI_ISL_423095\|20 | .................... | .................... | .................... | .................... | .................... | .................... | |
| A | 955 hCoV-19/England/20134017904/2020\|EPI_ISL_423096\|20 | .................... | .................... | .................... | .................... | .................... | .................... | |
| A | 956 hCoV-19/England/20134018004/2020\|EPI_ISL_423097\|20 | .................... | .................... | .................... | .................... | .................... | .................... | |
| A | 958 hCoV-19/England/20134018602/2020\|EPI_ISL_423099\|20 | .................... | .................... | .................... | .................... | .................... | .................... | |
| A | 959 hCoV-19/USA/FL-UFL-UF1/2020\|EPI_ISL_424350\|2020-03 | .................... | .................... | .................... | .................... | .................... | .................... | |
| A | 960 hCoV-19/USA/FL-UFL-UF2/2020\|EPI_ISL_424351\|2020-03 | .................... | .................... | .................... | .................... | .................... | .................... | |
| A | 961 hCoV-19/Fuyang/FY002/2020\|EPI_ISL_424352\|2020-03-1 | .................... | .................... | .................... | .................... | .................... | .................... | |
| A | 962 hCoV-19/USA/NC_200173/2020\|EPI_ISL_424353\|2020-03 | .................... | .................... | .................... | .................... | .................... | .................... | |
| A | 963 hCoV-19/USA/NC_200189/2020\|EPI_ISL_424354\|2020-03 | .................... | .................... | .................... | .................... | .................... | .................... | |
| A | 964 hCoV-19/Beijing/Wuhan_IME-BJ01/2020\|EPI_ISL_424355 | .................... | .................... | .................... | .................... | .................... | .................... | |
| A | 965 hCoV-19/Beijing/Wuhan_IME-BJ02/2020\|EPI_ISL_424356 | .................... | .................... | .................... | .................... | .................... | .................... | |
| A | 966 hCoV-19/Beijing/Wuhan_IME-BJ03/2020\|EPI_ISL_424357 | .................... | .................... | .................... | .................... | .................... | .................... | |
| A | 967 hCoV-19/Beijing/Wuhan_IME-BJ04/2020\|EPI_ISL_424358 | .................... | .................... | .................... | .................... | .................... | .................... | |
| A | 968 hCoV-19/Beijing/Wuhan_IME-BJ05/2020\|EPI_ISL_424359 | .................... | .................... | .................... | .................... | .................... | .................... | |
| A | 969 hCoV-19/Scotland/CVR139/2020\|EPI_ISL_425682\|2020-0 | .................... | .................... | .................... | .................... | .................... | .................... | |
| A | 970 hCoV-19/Scotland/CVR140/2020\|EPI_ISL_425684\|2020-0 | .................... | .................... | .................... | .................... | .................... | .................... | |
| A | 971 hCoV-19/Scotland/CVR141/2020\|EPI_ISL_425685\|2020-0 | .................... | .................... | .................... | .................... | .................... | .................... | |
| A | 972 hCoV-19/Scotland/CVR143/2020\|EPI_ISL_425687\|2020-0 | .................... | .................... | .................... | .................... | .................... | .................... | |
| A | 973 hCoV-19/Scotland/CVR144/2020\|EPI_ISL_425688\|2020-0 | .................... | .................... | .................... | .................... | .................... | .................... | |
| A | 974 hCoV-19/Scotland/CVR161/2020\|EPI_ISL_425700\|2020-0 | .................... | .A.................. | .................... | .................... | .................... | .................... | |
| A | 975 hCoV-19/Scotland/CVR162/2020\|EPI_ISL_425701\|2020-0 | .................... | .................... | .................... | .................... | .................... | .................... | |
| A | 976 hCoV-19/Switzerland/GE-HUG-5737/2020\|EPI_ISL_42920 | .................... | .................... | .................... | .................... | .................... | .................... | |
| A | 977 hCoV-19/Switzerland/GE-HUG-2297/2020\|EPI_ISL_42920 | .................... | .................... | .................... | .................... | .................... | .................... | |
| A | 978 hCoV-19/Switzerland/GE-HUG-1736/2020\|EPI_ISL_42920 | .................... | .................... | .................... | .................... | .................... | .................... | |
| A | 979 hCoV-19/Switzerland/GE-HUG-7617/2020\|EPI_ISL_42920 | .................... | .................... | .................... | .................... | .................... | .................... | |
| A | 980 hCoV-19/Switzerland/GE-HUG-5007/2020\|EPI_ISL_42920 | .................... | .................... | .................... | .................... | .................... | .................... | |
| A | 981 hCoV-19/Switzerland/GE-HUG-1184/2020\|EPI_ISL_42920 | .................... | .................... | .................... | .................... | .................... | .................... | |
| A | 982 hCoV-19/Switzerland/GE-HUG-2164/2020\|EPI_ISL_42920 | .................... | .................... | .................... | .................... | .................... | .................... | |
| A | 983 hCoV-19/Switzerland/GE-HUG-3650/2020\|EPI_ISL_42920 | .................... | ...................A | .................... | .................... | .................... | .................... | 1 |
| A | 984 hCoV-19/Switzerland/GE-HUG-8147/2020\|EPI_ISL_42921 | .................... | .A.................. | .................... | .................... | .................... | .................... | |
| A | 985 hCoV-19/Switzerland/GE-HUG-5708/2020\|EPI_ISL_42921 | .................... | .................... | .................... | .................... | .................... | .................... | |
| A | 986 hCoV-19/Switzerland/GE-HUG-0056/2020\|EPI_ISL_42921 | .................... | .................... | .................... | .................... | .................... | .................... | |
| A | 987 hCoV-19/Switzerland/GE-HUG-8086/2020\|EPI_ISL_42921 | .................... | .................... | .................... | .................... | .................... | .................... | |

| | | | | | | | | |
|---|---|---|---|---|---|---|---|---|
| A | 988 | hCoV-19/Switzerland/GE-HUG-2759/2020\|EPI_ISL_42921 | ···················· | ···················· | ···················· | ···················· | ···················· | ···················· |
| A | 989 | hCoV-19/Switzerland/GE-HUG-4807/2020\|EPI_ISL_42921 | ···················· | ···················· | ···················· | ···················· | ···················· | ···················· |
| A | 990 | hCoV-19/Switzerland/GE-HUG-6414/2020\|EPI_ISL_42921 | ···················· | ···················· | ···················· | ···················· | ···················· | ···················· |
| A | 991 | hCoV-19/Switzerland/GE-HUG-0304/2020\|EPI_ISL_42921 | ···················· | ·A·················· | ···················· | ···················· | ···················· | ···················· |
| A | 992 | hCoV-19/Switzerland/GE-HUG-2453/2020\|EPI_ISL_42921 | ···················· | ·A·················· | ···················· | ···················· | ···················· | ···················· |
| A | 993 | hCoV-19/Switzerland/GE-HUG-6065/2020\|EPI_ISL_42922 | ···················· | ···················· | ···················· | ···················· | ···················· | ···················· |
| A | 994 | hCoV-19/Switzerland/GE-HUG-6099/2020\|EPI_ISL_42922 | ···················· | ···················· | ···················· | ···················· | ···················· | ···················· |
| A | 995 | hCoV-19/Switzerland/GE-HUG-3655/2020\|EPI_ISL_42922 | ···················· | ···················· | ···················· | ···················· | ···················· | ···················· |
| A | 996 | hCoV-19/Switzerland/GE-HUG-4644/2020\|EPI_ISL_42922 | ···················· | ···················· | ···················· | ···················· | ···················· | ···················· |
| A | 997 | hCoV-19/Italy/ABR-IZSGC-TE5551/2020\|EPI_ISL_429226 | ···················· | ···················· | ···················· | ···················· | ···················· | ···················· |
| A | 998 | hCoV-19/Italy/ABR-IZSGC-TE5553/2020\|EPI_ISL_429227 | ···················· | ···················· | ···················· | ···················· | ···················· | ···················· |
| A | 999 | hCoV-19/Italy/ABR-IZSGC-TE5674/2020\|EPI_ISL_429228 | ···················· | ···················· | ···················· | ···················· | ···················· | ···················· |
| A | 1000 | hCoV-19/Italy/ABR-IZSGC-TE6572/2020\|EPI_ISL_429232 | ···················· | ···················· | ···················· | ···················· | ···················· | ···················· |
| A | 1001 | hCoV-19/Italy/ABR-IZSGC-TE6580/2020\|EPI_ISL_429235 | ···················· | ···················· | ···················· | ···················· | ···················· | ···················· |
| A | 1002 | hCoV-19/Italy/ABR-AMVRC-TE7859/2020\|EPI_ISL_429236 | ···················· | ···················· | ···················· | ···················· | ···················· | ···················· |
| A | 1003 | hCoV-19/Yunnan/0306-466/2020\|EPI_ISL_429239\|2020-0 | ···················· | ···················· | ···················· | ···················· | ···················· | ···················· |
| A | 1004 | hCoV-19/Denmark/ALAB-HH11/2020\|EPI_ISL_429270\|2020 | ···················· | ···················· | ···················· | ···················· | ···················· | ···················· |
| A | 1005 | hCoV-19/Denmark/ALAB-HH13/2020\|EPI_ISL_429271\|2020 | ···················· | ···················· | ···················· | ···················· | ···················· | ···················· |
| A | 1006 | hCoV-19/Denmark/ALAB-HH14/2020\|EPI_ISL_429272\|2020 | ···················· | ···················· | ···················· | ···················· | ···················· | ···················· |
| A | 1007 | hCoV-19/Denmark/ALAB-HH15/2020\|EPI_ISL_429273\|2020 | ···················· | ···················· | ···················· | ···················· | ···················· | ···················· |
| A | 1008 | hCoV-19/Denmark/ALAB-HH16/2020\|EPI_ISL_429274\|2020 | ···················· | ···················· | ···················· | ···················· | ···················· | ···················· |
| A | 1009 | hCoV-19/Denmark/ALAB-HH17/2020\|EPI_ISL_429275\|2020 | ···················· | ···················· | ···················· | ···················· | ···················· | ···················· |
| A | 1010 | hCoV-19/Denmark/ALAB-HH18/2020\|EPI_ISL_429276\|2020 | ···················· | ···················· | ···················· | ···················· | ···················· | ···················· |
| A | 1011 | hCoV-19/Denmark/ALAB-HH19/2020\|EPI_ISL_429277\|2020 | ···················· | ···················· | ···················· | ···················· | ···················· | ···················· |
| A | 1012 | hCoV-19/Denmark/ALAB-HH20/2020\|EPI_ISL_429278\|2020 | ···················· | ···················· | ···················· | ···················· | ···················· | ···················· |
| A | 1013 | hCoV-19/Denmark/ALAB-HH21/2020\|EPI_ISL_429279\|2020 | ···················· | ···················· | ···················· | ···················· | ···················· | ···················· |
| A | 1014 | hCoV-19/Denmark/ALAB-HH22/2020\|EPI_ISL_429280\|2020 | ···················· | ···················· | ···················· | ···················· | ···················· | ···················· |
| A | 1015 | hCoV-19/Denmark/ALAB-HH23/2020\|EPI_ISL_429281\|2020 | ···················· | ···················· | ···················· | ···················· | ···················· | ···················· |
| A | 1016 | hCoV-19/Denmark/ALAB-HH24/2020\|EPI_ISL_429282\|2020 | ···················· | ···················· | ···················· | ···················· | ···················· | ···················· |
| A | 1017 | hCoV-19/Denmark/ALAB-HH25/2020\|EPI_ISL_429283\|2020 | ···················· | ···················· | ···················· | ···················· | ···················· | ···················· |
| A | 1018 | hCoV-19/Denmark/ALAB-HH26/2020\|EPI_ISL_429284\|2020 | ···················· | ···················· | ···················· | ···················· | ···················· | ···················· |
| A | 1019 | hCoV-19/Denmark/ALAB-HH27/2020\|EPI_ISL_429285\|2020 | ···················· | ···················· | ···················· | ···················· | ···················· | ···················· |
| A | 1020 | hCoV-19/Denmark/ALAB-HH29/2020\|EPI_ISL_429287\|2020 | ···················· | ···················· | ···················· | ···················· | ···················· | ···················· |
| A | 1021 | hCoV-19/Denmark/ALAB-HH31/2020\|EPI_ISL_429289\|2020 | ···················· | ···················· | ···················· | ···················· | ···················· | ···················· |
| A | 1022 | hCoV-19/Denmark/ALAB-HH37/2020\|EPI_ISL_429293\|2020 | ···················· | ···················· | ···················· | ···················· | ···················· | ···················· |
| A | 1023 | hCoV-19/Denmark/ALAB-HH39/2020\|EPI_ISL_429295\|2020 | ···················· | ···················· | ···················· | ···················· | ···················· | ···················· |
| A | 1024 | hCoV-19/Denmark/ALAB-HH43/2020\|EPI_ISL_429299\|2020 | ···················· | ···················· | ···················· | ···················· | ···················· | ···················· |
| A | 1025 | hCoV-19/Fujian/8/2020\|EPI_ISL_411060\|2020-01-21 | ···················· | ···················· | ···················· | ···················· | ···················· | ···················· |
| A | 1026 | hCoV-19/Fujian/13/2020\|EPI_ISL_411066\|2020-01-22 | ···················· | ···················· | ···················· | ···················· | ···················· | ···················· |
| A | 1027 | hCoV-19/Beijing/ICDC-DT005/2020\|EPI_ISL_412386\|202 | ···················· | ···················· | ···················· | ···················· | ···················· | ···················· |
| A | 1028 | hCoV-19/Finland/FIN03032020A/2020\|EPI_ISL_413602\|2 | ···················· | ···················· | ···················· | ···················· | ···················· | ···················· |
| A | 1029 | hCoV-19/Finland/FIN03032020B/2020\|EPI_ISL_413603\|2 | ···················· | ···················· | ···················· | ···················· | ···················· | ···················· |
| A | 1030 | hCoV-19/Finland/FIN03032020C/2020\|EPI_ISL_413604\|2 | ···················· | ···················· | ···················· | ···················· | ···················· | ···················· |
| A | 1031 | hCoV-19/USA/CruiseA-1/2020\|EPI_ISL_413606\|2020-02- | ···················· | ···················· | ···················· | ···················· | ···················· | ···················· |
| A | 1032 | hCoV-19/USA/CruiseA-2/2020\|EPI_ISL_413607\|2020-02- | ···················· | ···················· | ···················· | ···················· | ···················· | ···················· |
| A | 1033 | hCoV-19/USA/CruiseA-3/2020\|EPI_ISL_413608\|2020-02- | ···················· | ···················· | ···················· | ···················· | ···················· | ···················· |
| A | 1034 | hCoV-19/USA/CruiseA-4/2020\|EPI_ISL_413609\|2020-02- | ···················· | ···················· | ···················· | ···················· | ···················· | ···················· |
| A | 1035 | hCoV-19/USA/CruiseA-5/2020\|EPI_ISL_413610\|2020-02- | ···················· | ···················· | ···················· | ···················· | ···················· | ···················· |
| A | 1036 | hCoV-19/USA/CruiseA-6/2020\|EPI_ISL_413611\|2020-02- | ···················· | ···················· | ···················· | ···················· | ···················· | ···················· |
| A | 1037 | hCoV-19/USA/CruiseA-7/2020\|EPI_ISL_413612\|2020-02- | ···················· | ···················· | ···················· | ···················· | ···················· | ···················· |
| A | 1038 | hCoV-19/USA/CruiseA-8/2020\|EPI_ISL_413613\|2020-02- | ···················· | ···················· | ···················· | ···················· | ···················· | ···················· |
| A | 1039 | hCoV-19/USA/CruiseA-9/2020\|EPI_ISL_413614\|2020-02- | ···················· | ···················· | ···················· | ···················· | ···················· | ···················· |
| A | 1040 | hCoV-19/USA/CruiseA-10/2020\|EPI_ISL_413615\|2020-02 | ···················· | ···················· | ···················· | ···················· | ···················· | ···················· |
| A | 1041 | hCoV-19/USA/CruiseA-11/2020\|EPI_ISL_413616\|2020-02 | ···················· | ···················· | ···················· | ···················· | ···················· | ···················· |
| A | 1042 | hCoV-19/USA/CruiseA-12/2020\|EPI_ISL_413617\|2020-02 | ···················· | ···················· | ···················· | ···················· | ···················· | ···················· |
| A | 1043 | hCoV-19/USA/CruiseA-13/2020\|EPI_ISL_413618\|2020-02 | ···················· | ···················· | ···················· | ···················· | ···················· | ···················· |
| A | 1044 | hCoV-19/USA/CruiseA-14/2020\|EPI_ISL_413619\|2020-02 | ···················· | ···················· | ···················· | ···················· | ···················· | ···················· |
| A | 1045 | hCoV-19/USA/CruiseA-15/2020\|EPI_ISL_413620\|2020-02 | ···················· | ···················· | ···················· | ···················· | ···················· | ···················· |
| A | 1046 | hCoV-19/USA/CruiseA-16/2020\|EPI_ISL_413621\|2020-02 | ···················· | ···················· | ···················· | ···················· | ···················· | ···················· |
| A | 1047 | hCoV-19/USA/CruiseA-17/2020\|EPI_ISL_413622\|2020-02 | ···················· | ···················· | ···················· | ···················· | ···················· | ···················· |
| A | 1048 | hCoV-19/USA/CruiseA-18/2020\|EPI_ISL_413623\|2020-02 | ···················· | ···················· | ···················· | ···················· | ···················· | ···················· |
| A | 1049 | hCoV-19/Weifang/WF0001/2020\|EPI_ISL_413691\|2020-01 | ···················· | ···················· | ···················· | ···················· | ···················· | ···················· |
| A | 1050 | hCoV-19/Weifang/WF0002/2020\|EPI_ISL_413692\|2020-01 | ···················· | ···················· | ···················· | ···················· | ···················· | ···················· |
| A | 1051 | hCoV-19/Weifang/WF0003/2020\|EPI_ISL_413693\|2020-01 | ···················· | ···················· | ···················· | ···················· | ···················· | ···················· |
| A | 1052 | hCoV-19/Weifang/WF0004/2020\|EPI_ISL_413694\|2020-01 | ···················· | ···················· | ···················· | ···················· | ···················· | ···················· |
| A | 1053 | hCoV-19/Weifang/WF0012/2020\|EPI_ISL_413697\|2020-02 | ···················· | ···················· | ···················· | ···················· | ···················· | ···················· |
| A | 1054 | hCoV-19/Shandong/LY003-2/2020\|EPI_ISL_414936\|2020- | ···················· | ···················· | ···················· | ···················· | ···················· | ···················· |
| A | 1055 | hCoV-19/Shandong/LY004-2/2020\|EPI_ISL_414937\|2020- | ···················· | ···················· | ···················· | ···················· | ···················· | ···················· |
| A | 1056 | hCoV-19/Shandong/LY005-2/2020\|EPI_ISL_414938\|2020- | ···················· | ···················· | ···················· | ···················· | ···················· | ···················· |
| A | 1057 | hCoV-19/Shandong/LY006-2/2020\|EPI_ISL_414939\|2020- | ···················· | ···················· | ···················· | ···················· | ···················· | ···················· |
| A | 1058 | hCoV-19/Shandong/LY007-2/2020\|EPI_ISL_414940\|2020- | ···················· | ···················· | ···················· | ···················· | ···················· | ···················· |
| A | 1059 | hCoV-19/Shandong/LY008-2/2020\|EPI_ISL_414941\|2020- | ···················· | ···················· | ···················· | ···················· | ···················· | ···················· |
| A | 1060 | hCoV-19/Australia/VIC89/2020\|EPI_ISL_419800\|2020-0 | ···················· | ···················· | ···················· | ···················· | ···················· | ···················· |
| A | 1061 | hCoV-19/Australia/VIC90/2020\|EPI_ISL_419801\|2020-0 | ···················· | ···················· | ···················· | ···················· | ···················· | ···················· |
| A | 1062 | hCoV-19/Australia/VIC45/2020\|EPI_ISL_419802\|2020-0 | ···················· | ···················· | ···················· | ···················· | ···················· | ···················· |
| A | 1063 | hCoV-19/Australia/VIC84/2020\|EPI_ISL_419803\|2020-0 | ···················· | ···················· | ···················· | ···················· | ···················· | ···················· |
| A | 1064 | hCoV-19/Australia/VIC91/2020\|EPI_ISL_419804\|2020-0 | ···················· | ·A·················· | ···················· | ···················· | ···················· | ···················· |
| A | 1065 | hCoV-19/Australia/VIC94/2020\|EPI_ISL_419805\|2020-0 | ···················· | ···················· | ···················· | ···················· | ···················· | ···················· |

| | | | | | | | | |
|---|---|---|---|---|---|---|---|---|
| A | 1066 | hCoV-19/Australia/VIC95/2020\|EPI_ISL_419806\|2020-0 | ···················· | ···················· | ···················· | ···················· | ···················· | ···················· |
| A | 1067 | hCoV-19/Australia/VIC96/2020\|EPI_ISL_419807\|2020-0 | ···················· | ···················· | ···················· | ···················· | ···················· | ···················· |
| A | 1068 | hCoV-19/Australia/VIC97/2020\|EPI_ISL_419808\|2020-0 | ···················· | ···················· | ···················· | ···················· | ···················· | ···················· |
| A | 1069 | hCoV-19/Australia/VIC92/2020\|EPI_ISL_419809\|2020-0 | ···················· | ···················· | ···················· | ···················· | ···················· | ···················· |
| A | 1070 | hCoV-19/Australia/VIC98/2020\|EPI_ISL_419810\|2020-0 | ···················· | ···················· | ···················· | ···················· | ···················· | ···················· |
| A | 1071 | hCoV-19/Australia/VIC99/2020\|EPI_ISL_419811\|2020-0 | ···················· | ···················· | ···················· | ···················· | ···················· | ···················· |
| A | 1072 | hCoV-19/Australia/VIC100/2020\|EPI_ISL_419812\|2020- | ···················· | ···················· | ···················· | ···················· | ···················· | ···················· |
| A | 1073 | hCoV-19/Australia/VIC101/2020\|EPI_ISL_419813\|2020- | ···················· | ···················· | ···················· | ···················· | ···················· | ···················· |
| A | 1074 | hCoV-19/Australia/VIC103/2020\|EPI_ISL_419814\|2020- | ···················· | ···················· | ···················· | ···················· | ···················· | ···················· |
| A | 1075 | hCoV-19/Australia/VIC46/2020\|EPI_ISL_419816\|2020-0 | ···················· | ···················· | ···················· | ···················· | ···················· | ···················· |
| A | 1076 | hCoV-19/Australia/VIC102/2020\|EPI_ISL_419817\|2020- | ···················· | ···················· | ···················· | ···················· | ···················· | ···················· |
| A | 1077 | hCoV-19/Australia/VIC104/2020\|EPI_ISL_419818\|2020- | ···················· | ···················· | ···················· | ···················· | ···················· | ···················· |
| A | 1078 | hCoV-19/Australia/VIC105/2020\|EPI_ISL_419819\|2020- | ···················· | ···················· | ···················· | ···················· | ···················· | ···················· |
| A | 1079 | hCoV-19/Australia/VIC106/2020\|EPI_ISL_419820\|2020- | ···················· | ·A·················· | ···················· | ···················· | ···················· | ···················· |
| A | 1080 | hCoV-19/Australia/VIC112/2020\|EPI_ISL_419821\|2020- | ···················· | ···················· | ···················· | ···················· | ···················· | ···················· |
| A | 1081 | hCoV-19/Australia/VIC113/2020\|EPI_ISL_419822\|2020- | ···················· | ···················· | ···················· | ···················· | ···················· | ···················· |
| A | 1082 | hCoV-19/Australia/VIC130/2020\|EPI_ISL_419823\|2020- | ···················· | ···················· | ···················· | ···················· | ···················· | ···················· |
| A | 1083 | hCoV-19/Australia/VIC131/2020\|EPI_ISL_419824\|2020- | ···················· | ···················· | ···················· | ···················· | ···················· | ···················· |
| A | 1084 | hCoV-19/Australia/VIC132/2020\|EPI_ISL_419825\|2020- | ···················· | ···················· | ···················· | ···················· | ···················· | ···················· |
| A | 1085 | hCoV-19/Australia/VIC51/2020\|EPI_ISL_419826\|2020-0 | ···················· | ···················· | ···················· | ···················· | ···················· | ···················· |
| A | 1086 | hCoV-19/Australia/VIC134/2020\|EPI_ISL_419827\|2020- | ···················· | ···················· | ···················· | ···················· | ···················· | ···················· |
| A | 1087 | hCoV-19/South Korea/BA-ACH_2718/2020\|EPI_ISL_42080 | ···················· | ···················· | ···················· | ···················· | ···················· | ···················· |
| A | 1088 | hCoV-19/South Korea/BA-ACH_2719/2020\|EPI_ISL_42080 | ···················· | ···················· | ···················· | ···················· | ···················· | ···················· |
| A | 1089 | hCoV-19/USA/UT-089/2020\|EPI_ISL_420803\|2020-03-12 | ···················· | ···················· | ···················· | ···················· | ···················· | ···················· |
| A | 1090 | hCoV-19/USA/UT-0289/2020\|EPI_ISL_420809\|2020-03-24 | ···················· | ···················· | ···················· | ···················· | ···················· | ···················· |
| A | 1091 | hCoV-19/USA/WA-UW-1448/2020\|EPI_ISL_423007\|2020-03 | ···················· | ···················· | ···················· | ···················· | ···················· | ···················· |
| A | 1092 | hCoV-19/USA/WA-UW-1426/2020\|EPI_ISL_423008\|2020-03 | ···················· | ···················· | ···················· | ···················· | ···················· | ···················· |
| A | 1093 | hCoV-19/USA/WA-UW-1446/2020\|EPI_ISL_423010\|2020-03 | ···················· | ···················· | ···················· | ···················· | ···················· | ···················· |
| A | 1094 | hCoV-19/USA/WA-UW-1561/2020\|EPI_ISL_423011\|2020-03 | ···················· | ···················· | ···················· | ···················· | ···················· | ···················· |
| A | 1095 | hCoV-19/USA/WA-UW-1588/2020\|EPI_ISL_423012\|2020-03 | ···················· | ···················· | ···················· | ···················· | ···················· | ···················· |
| A | 1096 | hCoV-19/USA/WA-UW-1586/2020\|EPI_ISL_423013\|2020-03 | ···················· | ···················· | ···················· | ···················· | ···················· | ···················· |
| A | 1097 | hCoV-19/USA/WA-UW-1587/2020\|EPI_ISL_423014\|2020-03 | ···················· | ···················· | ···················· | ···················· | ···················· | ···················· |
| A | 1098 | hCoV-19/USA/WA-UW-1552/2020\|EPI_ISL_423015\|2020-03 | ···················· | ···················· | ···················· | ···················· | ···················· | ···················· |
| A | 1099 | hCoV-19/USA/WA-UW-1578/2020\|EPI_ISL_423016\|2020-03 | ···················· | ···················· | ···················· | ···················· | ···················· | ···················· |
| A | 1100 | hCoV-19/USA/WA-UW-1591/2020\|EPI_ISL_423017\|2020-03 | ···················· | ···················· | ···················· | ···················· | ···················· | ···················· |
| A | 1101 | hCoV-19/USA/WA-UW-1576/2020\|EPI_ISL_423018\|2020-03 | ···················· | ···················· | ···················· | ···················· | ···················· | ···················· |
| A | 1102 | hCoV-19/USA/WA-UW-1574/2020\|EPI_ISL_423019\|2020-03 | ···················· | ···················· | ···················· | ···················· | ···················· | ···················· |
| A | 1103 | hCoV-19/USA/WA-UW-1570/2020\|EPI_ISL_423020\|2020-03 | ···················· | ···················· | ···················· | ···················· | ···················· | ···················· |
| A | 1104 | hCoV-19/USA/WA-UW-1471/2020\|EPI_ISL_423029\|2020-03 | ···················· | ···················· | ···················· | ···················· | ···················· | ···················· |
| A | 1105 | hCoV-19/USA/WA-UW-1433/2020\|EPI_ISL_423030\|2020-03 | ···················· | ···················· | ···················· | ···················· | ···················· | ···················· |
| A | 1106 | hCoV-19/USA/WA-UW-1425/2020\|EPI_ISL_423031\|2020-03 | ···················· | ···················· | ···················· | ···················· | ···················· | ···················· |
| A | 1107 | hCoV-19/USA/WA-UW-1608/2020\|EPI_ISL_423032\|2020-03 | ···················· | ···················· | ···················· | ···················· | ···················· | ···················· |
| A | 1108 | hCoV-19/USA/WA-UW-1599/2020\|EPI_ISL_423033\|2020-03 | ···················· | ···················· | ···················· | ···················· | ···················· | ···················· |
| A | 1111 | hCoV-19/Thailand/Bangkok-0020/2020\|EPI_ISL_423041\| | ···················· | ···················· | ···················· | ···················· | ···················· | ···················· |
| A | 1112 | hCoV-19/Thailand/Bangkok-0021/2020\|EPI_ISL_423042\| | ···················· | ···················· | ···················· | ···················· | ···················· | ···················· |
| A | 1113 | hCoV-19/Thailand/Bangkok-0022/2020\|EPI_ISL_423043\| | ···················· | ···················· | ···················· | ···················· | ···················· | ···················· |
| A | 1114 | hCoV-19/England/20134000704/2020\|EPI_ISL_423044\|20 | ···················· | ···················· | ···················· | ···················· | ···················· | ···················· |
| A | 1115 | hCoV-19/England/20134000804/2020\|EPI_ISL_423045\|20 | ···················· | ···················· | ···················· | ···················· | ···················· | ···················· |
| A | 1116 | hCoV-19/England/20134001104/2020\|EPI_ISL_423047\|20 | ···················· | ···················· | ···················· | ···················· | ···················· | ···················· |
| A | 1117 | hCoV-19/England/20134001904/2020\|EPI_ISL_423052\|20 | ···················· | ···················· | ···················· | ···················· | ···················· | ···················· |
| A | 1118 | hCoV-19/England/20134002104/2020\|EPI_ISL_423053\|20 | ···················· | ···················· | ···················· | ···················· | ···················· | ···················· |
| A | 1121 | hCoV-19/England/20134003204/2020\|EPI_ISL_423057\|20 | ···················· | ···················· | ···················· | ···················· | ···················· | ···················· |
| A | 1122 | hCoV-19/England/20134003304/2020\|EPI_ISL_423058\|20 | ···················· | ···················· | ···················· | ···················· | ···················· | ···················· |
| A | 1124 | hCoV-19/England/20134004704/2020\|EPI_ISL_423063\|20 | ···················· | ···················· | ···················· | ···················· | ···················· | ···················· |
| A | 1125 | hCoV-19/England/20134004904/2020\|EPI_ISL_423065\|20 | ···················· | ···················· | ···················· | ···················· | ···················· | ···················· |
| A | 1126 | hCoV-19/England/20134005004/2020\|EPI_ISL_423066\|20 | ···················· | ···················· | ···················· | ···················· | ···················· | ···················· |
| A | 1127 | hCoV-19/England/20134005104/2020\|EPI_ISL_423067\|20 | ···················· | ···················· | ···················· | ···················· | ···················· | ···················· |
| A | 1128 | hCoV-19/England/20134005304/2020\|EPI_ISL_423068\|20 | ···················· | ···················· | ···················· | ···················· | ···················· | ···················· |
| A | 1130 | hCoV-19/England/20134006504/2020\|EPI_ISL_423070\|20 | ···················· | ···················· | ···················· | ···················· | ···················· | ···················· |
| A | 1131 | hCoV-19/England/20134006604/2020\|EPI_ISL_423071\|20 | ···················· | ···················· | ···················· | ···················· | ···················· | ···················· |
| A | 1132 | hCoV-19/England/20134006704/2020\|EPI_ISL_423072\|20 | ···················· | ···················· | ···················· | ···················· | ···················· | ···················· |
| A | 1133 | hCoV-19/England/20134007004/2020\|EPI_ISL_423073\|20 | ···················· | ···················· | ···················· | ···················· | ···················· | ···················· |
| A | 1134 | hCoV-19/England/20134007104/2020\|EPI_ISL_423074\|20 | ···················· | ···················· | ···················· | ···················· | ···················· | ···················· |
| A | 1135 | hCoV-19/England/20134007204/2020\|EPI_ISL_423075\|20 | ···················· | ···················· | ···················· | ···················· | ···················· | ················N··· |
| A | 1136 | hCoV-19/England/20134009504/2020\|EPI_ISL_423076\|20 | ···················· | ···················· | ···················· | ···················· | ···················· | ···················· |
| A | 1137 | hCoV-19/England/20134010904/2020\|EPI_ISL_423078\|20 | ···················· | ···················· | ···················· | ···················· | ···················· | ···················· |
| A | 1138 | hCoV-19/England/20134011704/2020\|EPI_ISL_423079\|20 | ···················· | ···················· | ···················· | ···················· | ···················· | ···················· |
| A | 1139 | hCoV-19/England/201340120/2020\|EPI_ISL_423081\|2020 | ···················· | ···················· | ···················· | ···················· | ···················· | ···················· |
| A | 1140 | hCoV-19/England/201340127/2020\|EPI_ISL_423084\|2020 | ···················· | ···················· | ···················· | ···················· | ···················· | ···················· |
| A | 1141 | hCoV-19/England/20134015504/2020\|EPI_ISL_423085\|20 | ···················· | ···················· | ···················· | ···················· | ··N················· | ···················· |
| A | 1142 | hCoV-19/England/20134015704/2020\|EPI_ISL_423086\|20 | ···················· | ···················· | ···················· | ···················· | ···················· | ···················· |
| A | 1144 | hCoV-19/USA/WA-UW-1878/2020\|EPI_ISL_424302\|2020-03 | ···················· | ···················· | ···················· | ···················· | ···················· | ···················· |
| A | 1145 | hCoV-19/USA/WA-UW-1930/2020\|EPI_ISL_424304\|2020-03 | ···················· | ···················· | ···················· | ···················· | ···················· | ···················· |
| A | 1146 | hCoV-19/USA/WA-UW-1877/2020\|EPI_ISL_424305\|2020-03 | ···················· | ···················· | ···················· | ···················· | ···················· | ···················· |
| A | 1147 | hCoV-19/USA/WA-UW-1874/2020\|EPI_ISL_424306\|2020-03 | ···················· | ···················· | ···················· | ···················· | ···················· | ···················· |
| A | 1148 | hCoV-19/USA/OR-UW-1885/2020\|EPI_ISL_424307\|2020-03 | ···················· | ···················· | ···················· | ···················· | ···················· | ···················· |
| A | 1149 | hCoV-19/USA/OR-UW-1887/2020\|EPI_ISL_424308\|2020-03 | ···················· | ···················· | ···················· | ···················· | ···················· | ···················· |
| A | 1150 | hCoV-19/USA/WA-UW-1881/2020\|EPI_ISL_424309\|2020-03 | ···················· | ···················· | ···················· | ···················· | ···················· | ···················· |

| | | | | | | | | |
|---|---|---|---|---|---|---|---|---|
| A | 1151 | hCoV-19/USA/WA-UW-1888/2020\|EPI_ISL_424310\|2020-03 | .................... | .................... | .................... | .................... | .................... | .................... |
| A | 1152 | hCoV-19/USA/WA-UW-1880/2020\|EPI_ISL_424311\|2020-03 | .................... | .................... | .................... | .................... | .................... | .................... |
| A | 1153 | hCoV-19/USA/WA-UW-1955/2020\|EPI_ISL_424312\|2020-03 | .................... | .................... | .................... | .................... | .................... | .................... |
| A | 1154 | hCoV-19/USA/WA-UW-1956/2020\|EPI_ISL_424313\|2020-03 | .................... | .................... | .................... | .................... | .................... | .................... |
| A | 1155 | hCoV-19/USA/WA-UW-1921/2020\|EPI_ISL_424314\|2020-03 | .................... | .................... | .................... | .................... | .................... | .................... |
| A | 1156 | hCoV-19/USA/ID-UW-1957/2020\|EPI_ISL_424315\|2020-03 | .................... | .................... | .................... | .................... | .................... | .................... |
| A | 1157 | hCoV-19/USA/WA-UW-1986/2020\|EPI_ISL_424338\|2020-03 | .................... | .................... | .................... | .................... | .................... | .................... |
| A | 1158 | hCoV-19/USA/WA-UW-2016/2020\|EPI_ISL_424339\|2020-03 | .................... | .................... | .................... | .................... | .................... | .................... |
| A | 1159 | hCoV-19/USA/WA-UW-2009/2020\|EPI_ISL_424340\|2020-03 | .................... | .................... | .................... | .................... | .................... | .................... |
| A | 1160 | hCoV-19/USA/WA-UW-2008/2020\|EPI_ISL_424341\|2020-03 | .................... | .................... | .................... | .................... | .................... | .................... |
| A | 1161 | hCoV-19/Italy/LAZ-INMI-8/2020\|EPI_ISL_424342\|2020- | .................... | .................... | .................... | .................... | .................... | .................... |
| A | 1162 | hCoV-19/Italy/LAZ-INMI-9/2020\|EPI_ISL_424343\|2020- | .................... | .................... | .................... | .................... | .................... | .................... |
| A | 1163 | hCoV-19/Italy/LAZ-INMI10/2020\|EPI_ISL_424344\|2020- | .................... | .................... | .................... | .................... | .................... | .................... |
| A | 1164 | hCoV-19/Mexico/CMX-INER-01/2020\|EPI_ISL_424345\|202 | .................... | .................... | .................... | .................... | .................... | .................... |
| A | 1165 | hCoV-19/USA/AK-PHL037/2020\|EPI_ISL_424346\|2020-03- | .................... | .................... | .................... | .................... | .................... | .................... |
| A | 1166 | hCoV-19/USA/AK-PHL077/2020\|EPI_ISL_424347\|2020-03- | .................... | .................... | .................... | .................... | .................... | .................... |
| A | 1167 | hCoV-19/Beijing/Wuhan_IME-BJ07/2020\|EPI_ISL_424360 | .................... | .................... | .................... | .................... | .................... | .................... |
| A | 1168 | hCoV-19/India/UN-1063/2020\|EPI_ISL_424361\|2020-03- | .................... | .................... | .................... | .................... | .................... | .................... |
| A | 1169 | hCoV-19/India/UN-1135/2020\|EPI_ISL_424362\|2020-03- | .................... | .................... | .................... | .................... | .................... | .................... |
| A | 1170 | hCoV-19/India/UN-1652/2020\|EPI_ISL_424363\|2020-03- | .................... | .................... | .................... | .................... | .................... | .................... |
| A | 1171 | hCoV-19/India/UN-3118/2020\|EPI_ISL_424364\|2020-03- | .................... | .................... | .................... | .................... | .................... | .................... |
| A | 1172 | hCoV-19/India/UN-3239/2020\|EPI_ISL_424365\|2020-03- | .................... | .................... | .................... | .................... | .................... | .................... |
| A | 1173 | hCoV-19/Turkey/ERAGEM-001/2020\|EPI_ISL_424366\|2020 | .................... | .................... | .................... | .................... | .................... | .................... |
| A | 1174 | hCoV-19/Iceland/343/2020\|EPI_ISL_424367\|2020-03 | .................... | .................... | .................... | .................... | .................... | .................... |
| A | 1175 | hCoV-19/Iceland/344/2020\|EPI_ISL_424368\|2020-03 | .................... | .................... | .................... | .................... | .................... | .................... |
| A | 1176 | hCoV-19/Iceland/345/2020\|EPI_ISL_424369\|2020-03 | .................... | .................... | .................... | .................... | .................... | .................... |
| A | 1177 | hCoV-19/Iceland/346/2020\|EPI_ISL_424370\|2020-03 | .................... | .................... | .................... | .................... | .................... | .................... |
| A | 1178 | hCoV-19/Iceland/347/2020\|EPI_ISL_424371\|2020-03 | .................... | .................... | .................... | .................... | .................... | .................... |
| A | 1179 | hCoV-19/Iceland/351/2020\|EPI_ISL_424375\|2020-03 | .................... | .................... | .................... | .................... | .................... | .................... |
| A | 1180 | hCoV-19/Iceland/352/2020\|EPI_ISL_424376\|2020-03 | .................... | .................... | .................... | .................... | .................... | .................... |
| A | 1181 | hCoV-19/Iceland/353/2020\|EPI_ISL_424377\|2020-03-18 | .................... | .................... | .................... | .................... | .................... | .................... |
| A | 1182 | hCoV-19/Iceland/355/2020\|EPI_ISL_424379\|2020-03-19 | .................... | .................... | .................... | .................... | .................... | .................... |
| A | 1183 | hCoV-19/Iceland/356/2020\|EPI_ISL_424380\|2020-03-19 | .................... | .................... | .................... | .................... | .................... | .................... |
| A | 1184 | hCoV-19/Iceland/357/2020\|EPI_ISL_424381\|2020-03-19 | .................... | .................... | .................... | .................... | .................... | .................... |
| A | 1185 | hCoV-19/Iceland/358/2020\|EPI_ISL_424382\|2020-03-19 | .................... | .................... | .................... | .................... | .................... | .................... |
| A | 1186 | hCoV-19/Iceland/359/2020\|EPI_ISL_424383\|2020-03-19 | .................... | .................... | .................... | .................... | .................... | .................... |
| A | 1187 | hCoV-19/Iceland/360/2020\|EPI_ISL_424384\|2020-03-19 | .................... | .................... | .................... | .................... | .................... | .................... |
| A | 1188 | hCoV-19/Iceland/361/2020\|EPI_ISL_424385\|2020-03-19 | .................... | .................... | .................... | .................... | .................... | .................... |
| A | 1189 | hCoV-19/Iceland/363/2020\|EPI_ISL_424387\|2020-03-19 | .................... | .................... | .................... | .................... | .................... | .................... |
| A | 1190 | hCoV-19/Iceland/364/2020\|EPI_ISL_424388\|2020-03-19 | .................... | .................... | .................... | .................... | .................... | .................... |
| A | 1191 | hCoV-19/Iceland/365/2020\|EPI_ISL_424389\|2020-03-19 | .................... | .................... | .................... | .................... | .................... | .................... |
| A | 1192 | hCoV-19/Iceland/366/2020\|EPI_ISL_424390\|2020-03-19 | .................... | .................... | .................... | .................... | .................... | .................... |
| A | 1193 | hCoV-19/Iceland/367/2020\|EPI_ISL_424391\|2020-03-19 | .................... | .................... | .................... | .................... | .................... | .................... |
| A | 1194 | hCoV-19/Iceland/368/2020\|EPI_ISL_424392\|2020-03-19 | .................... | .................... | .................... | .................... | .................... | .................... |
| A | 1195 | hCoV-19/Iceland/369/2020\|EPI_ISL_424393\|2020-03-19 | .................... | .................... | .................... | .................... | .................... | .................... |
| A | 1196 | hCoV-19/Iceland/370/2020\|EPI_ISL_424394\|2020-03-19 | .................... | .................... | .................... | .................... | .................... | .................... |
| A | 1197 | hCoV-19/Iceland/371/2020\|EPI_ISL_424395\|2020-03-19 | .................... | .................... | .................... | .................... | .................... | .................... |
| A | 1198 | hCoV-19/Iceland/372/2020\|EPI_ISL_424396\|2020-03-19 | .................... | .................... | .................... | .................... | .................... | .................... |
| A | 1199 | hCoV-19/Iceland/374/2020\|EPI_ISL_424398\|2020-03-19 | .................... | .................... | .................... | .................... | .................... | .................... |
| A | 1200 | hCoV-19/Iceland/375/2020\|EPI_ISL_424399\|2020-03-19 | .................... | .................... | .................... | .................... | .................... | .................... |
| A | 1201 | hCoV-19/England/NOTT-10E836/2020\|EPI_ISL_425600\|20 | .................... | .................... | .................... | .................... | .................... | .................... |
| A | 1202 | hCoV-19/England/NOTT-10E845/2020\|EPI_ISL_425601\|20 | .................... | .................... | .................... | .................... | .................... | .................... |
| A | 1203 | hCoV-19/England/NOTT-10E872/2020\|EPI_ISL_425603\|20 | .................... | .................... | .................... | .................... | .................... | .................... |
| A | 1204 | hCoV-19/England/NOTT-10E8DC/2020\|EPI_ISL_425609\|20 | .................... | .................... | .................... | .................... | .................... | .................... |
| A | 1205 | hCoV-19/England/NOTT-10E8FA/2020\|EPI_ISL_425611\|20 | .................... | .................... | .................... | .................... | .................... | .................... |
| A | 1206 | hCoV-19/England/NOTT-10E933/2020\|EPI_ISL_425614\|20 | .................... | .................... | .................... | .................... | .................... | .................... |
| A | 1207 | hCoV-19/England/NOTT-10E942/2020\|EPI_ISL_425615\|20 | .................... | .................... | .................... | .................... | .................... | .................... |
| A | 1208 | hCoV-19/England/NOTT-10E951/2020\|EPI_ISL_425616\|20 | .................... | .................... | .................... | .................... | .................... | .................... |
| A | 1209 | hCoV-19/England/NOTT-10E97F/2020\|EPI_ISL_425618\|20 | .................... | .................... | .................... | .................... | .................... | .................... |
| A | 1210 | hCoV-19/England/NOTT-10E9AC/2020\|EPI_ISL_425620\|20 | .................... | .................... | .................... | .................... | .................... | .................... |
| A | 1211 | hCoV-19/England/NOTT-10E9BB/2020\|EPI_ISL_425621\|20 | .................... | .................... | .................... | .................... | .................... | .................... |
| A | 1212 | hCoV-19/England/NOTT-10E9CA/2020\|EPI_ISL_425622\|20 | .................... | .................... | .................... | .................... | .................... | .................... |
| A | 1213 | hCoV-19/England/NOTT-10E9D9/2020\|EPI_ISL_425623\|20 | .................... | .................... | .................... | .................... | .................... | .................... |
| A | 1214 | hCoV-19/England/NOTT-10EA8B/2020\|EPI_ISL_425633\|20 | .................... | .................... | .................... | .................... | .................... | .................... |
| A | 1215 | hCoV-19/England/NOTT-10EAA9/2020\|EPI_ISL_425635\|20 | .................... | .................... | .................... | .................... | .................... | .................... |
| A | 1216 | hCoV-19/England/NOTT-10EAC7/2020\|EPI_ISL_425637\|20 | .................... | .................... | .................... | .................... | .................... | .................... |
| A | 1217 | hCoV-19/England/NOTT-10EAD6/2020\|EPI_ISL_425638\|20 | .................... | .................... | .................... | .................... | .................... | .................... |
| A | 1218 | hCoV-19/England/NOTT-10EAE5/2020\|EPI_ISL_425639\|20 | .................... | .................... | .................... | .................... | .................... | .................... |
| A | 1219 | hCoV-19/England/NOTT-10EAF4/2020\|EPI_ISL_425640\|20 | .................... | .................... | .................... | .................... | .................... | .................... |
| A | 1220 | hCoV-19/England/NOTT-10EB1F/2020\|EPI_ISL_425641\|20 | .................... | .................... | .................... | .................... | .................... | .................... |
| A | 1221 | hCoV-19/England/NOTT-10EB2E/2020\|EPI_ISL_425642\|20 | .................... | .................... | .................... | .................... | .................... | .................... |
| A | 1222 | hCoV-19/England/NOTT-10EB3D/2020\|EPI_ISL_425643\|20 | .................... | .................... | .................... | .................... | .................... | .................... |
| A | 1223 | hCoV-19/England/NOTT-10EB6A/2020\|EPI_ISL_425644\|20 | .................... | .................... | .................... | .................... | .................... | .................... |
| A | 1224 | hCoV-19/England/NOTT-10EB79/2020\|EPI_ISL_425645\|20 | .................... | .................... | .................... | .................... | .................... | .................... |
| A | 1225 | hCoV-19/Scotland/CVR126/2020\|EPI_ISL_425669\|2020-0 | .................... | .................... | .................... | .................... | .................... | .................... |
| A | 1226 | hCoV-19/Scotland/CVR128/2020\|EPI_ISL_425670\|2020-0 | .................... | .................... | .................... | .................... | .................... | .................... |
| A | 1227 | hCoV-19/Scotland/CVR129/2020\|EPI_ISL_425671\|2020-0 | .................... | .................... | .................... | .................... | .................... | .................... |
| A | 1228 | hCoV-19/Scotland/CVR13/2020\|EPI_ISL_425672\|2020-03 | .................... | .................... | .................... | .................... | .................... | .................... |

| | | | | | | | |
|---|---|---|---|---|---|---|---|
| A | 1229 hCoV-19/Scotland/CVR131/2020\|EPI_ISL_425674\|2020-0 | .................... | .................... | .................... | .................... | .................... | .................... |
| A | 1230 hCoV-19/Scotland/CVR132/2020\|EPI_ISL_425675\|2020-0 | .................... | .................... | .................... | .................... | .................... | .................... |
| A | 1231 hCoV-19/Scotland/CVR133/2020\|EPI_ISL_425676\|2020-0 | .................... | .................... | .................... | .................... | .................... | .................... |
| A | 1232 hCoV-19/Scotland/CVR134/2020\|EPI_ISL_425677\|2020-0 | .................... | .................... | .................... | .................... | .................... | .................... |
| A | 1233 hCoV-19/Scotland/CVR135/2020\|EPI_ISL_425678\|2020-0 | .................... | .................... | .................... | .................... | .................... | .................... |
| A | 1234 hCoV-19/Scotland/CVR136/2020\|EPI_ISL_425679\|2020-0 | .................... | .................... | .................... | .................... | .................... | .................... |
| A | 1235 hCoV-19/Scotland/CVR15/2020\|EPI_ISL_425692\|2020-03 | .................... | .................... | .................... | .................... | .................... | .................... |
| A | 1236 hCoV-19/Scotland/CVR150/2020\|EPI_ISL_425693\|2020-0 | .................... | .................... | .................... | .................... | .................... | .................... |
| A | 1237 hCoV-19/Scotland/CVR152/2020\|EPI_ISL_425695\|2020-0 | .................... | .................... | .................... | .................... | .................... | .................... |
| A | 1238 hCoV-19/Scotland/CVR153/2020\|EPI_ISL_425696\|2020-0 | .................... | .................... | .................... | .................... | .................... | .................... |
| A | 1239 hCoV-19/Scotland/CVR155/2020\|EPI_ISL_425697\|2020-0 | .................... | .................... | .................... | .................... | .................... | .................... |
| A | 1240 hCoV-19/Scotland/CVR159/2020\|EPI_ISL_425698\|2020-0 | .................... | .................... | .................... | .................... | .................... | .................... |
| A | 1241 hCoV-19/Scotland/CVR16/2020\|EPI_ISL_425699\|2020-03 | .................... | .................... | .................... | .................... | .................... | .................... |
| A | 1252 hCoV-19/Australia/VIC623/2020\|EPI_ISL_426927\|2020- | .................... | .................... | .................... | .................... | .................... | .................... |
| A | 1256 hCoV-19/Australia/VIC628/2020\|EPI_ISL_426932\|2020- | .................... | .................... | .................... | .................... | .................... | .................... |
| A | 1258 hCoV-19/Australia/VIC638/2020\|EPI_ISL_426934\|2020- | .................... | .................... | .................... | .................... | .................... | .................... |
| A | 1259 hCoV-19/Australia/VIC639/2020\|EPI_ISL_426935\|2020- | .................... | .................... | .................... | .................... | .................... | .................... |
| A | 1260 hCoV-19/Australia/VIC644/2020\|EPI_ISL_426940\|2020- | .................... | .................... | .................... | .................... | .................... | .................... |
| A | 1261 hCoV-19/Australia/VIC645/2020\|EPI_ISL_426941\|2020- | .................... | .................... | .................... | .................... | .................... | .................... |
| A | 1262 hCoV-19/Australia/VIC646/2020\|EPI_ISL_426942\|2020- | .................... | .................... | .................... | .................... | .................... | .................... |
| A | 1263 hCoV-19/Australia/VIC647/2020\|EPI_ISL_426943\|2020- | .................... | .................... | .................... | .................... | .................... | .................... |
| A | 1265 hCoV-19/Australia/VIC652/2020\|EPI_ISL_426948\|2020- | .................... | .................... | .................... | .................... | .................... | .................... |
| A | 1267 hCoV-19/Australia/VIC656/2020\|EPI_ISL_426952\|2020- | .................... | .................... | .................... | .................... | .................... | .................... |
| A | 1272 hCoV-19/Australia/VIC685/2020\|EPI_ISL_426980\|2020- | .................... | .................... | .................... | .................... | .................... | .................... |
| A | 1275 hCoV-19/Australia/VIC694/2020\|EPI_ISL_426986\|2020- | .................... | .................... | .................... | .................... | .................... | .................... |
| A | 1276 hCoV-19/Australia/VIC696/2020\|EPI_ISL_426988\|2020- | .................... | .................... | .................... | .................... | .................... | .................... |
| A | 1277 hCoV-19/Australia/VIC697/2020\|EPI_ISL_426989\|2020- | .................... | .................... | .................... | .................... | .................... | .................... |
| A | 1278 hCoV-19/Switzerland/GE-HUG-3144/2020\|EPI_ISL_42919 | .................... | .................... | .................... | .................... | .................... | .................... |
| A | 1279 hCoV-19/Switzerland/GE-HUG-0199-2/2020\|EPI_ISL_429 | .................... | .A.................. | .................... | .................... | .................... | .................... |
| A | 1280 hCoV-19/Switzerland/GE-HUG-0636/2020\|EPI_ISL_42919 | .................... | .................... | .................... | .................... | .................... | .................... |
| A | 1281 hCoV-19/Portugal/CV62/2020\|EPI_ISL_413647\|2020-03- | .................... | .................... | .................... | .................... | .................... | .................... |
| A | 1282 hCoV-19/Portugal/CV63/2020\|EPI_ISL_413648\|2020-03- | .................... | .................... | .................... | .................... | .................... | .................... |
| A | 1283 hCoV-19/USA/WA14-UW10/2020\|EPI_ISL_413649\|2020-03- | .................... | .................... | .................... | .................... | .................... | .................... |
| A | 1284 hCoV-19/USA/WA15-UW11/2020\|EPI_ISL_413650\|2020-03- | .................... | .................... | .................... | .................... | .................... | .................... |
| A | 1285 hCoV-19/USA/WA16-UW12/2020\|EPI_ISL_413651\|2020-03- | .................... | .................... | .................... | .................... | .................... | .................... |
| A | 1286 hCoV-19/USA/WA17-UW13/2020\|EPI_ISL_413652\|2020-03- | .................... | .................... | .................... | .................... | .................... | .................... |
| A | 1287 hCoV-19/USA/WA18-UW14/2020\|EPI_ISL_413653\|2020-03- | .................... | .................... | .................... | .................... | .................... | .................... |
| A | 1288 hCoV-19/France/HDF-1645/2020\|EPI_ISL_418220\|2020-0 | .................... | .................... | .................... | .................... | .................... | .................... |
| A | 1289 hCoV-19/France/HDF-1813/2020\|EPI_ISL_418221\|2020-0 | .................... | .................... | .................... | .................... | .................... | .................... |
| A | 1290 hCoV-19/France/CVL-2000/2020\|EPI_ISL_418222\|2020-0 | .................... | .................... | .................... | .................... | .................... | .................... |
| A | 1291 hCoV-19/France/HDF-2060/2020\|EPI_ISL_418223\|2020-0 | .................... | .................... | .................... | .................... | .................... | .................... |
| A | 1292 hCoV-19/France/HDF-2150/2020\|EPI_ISL_418224\|2020-0 | .................... | .................... | .................... | .................... | .................... | .................... |
| A | 1293 hCoV-19/France/HDF-2155/2020\|EPI_ISL_418225\|2020-0 | .................... | .................... | .................... | .................... | .................... | .................... |
| A | 1294 hCoV-19/France/HDF-2381/2020\|EPI_ISL_418226\|2020-0 | .................... | .................... | .................... | .................... | .................... | .................... |
| A | 1295 hCoV-19/France/HDF-2393/2020\|EPI_ISL_418227\|2020-0 | .................... | .................... | .................... | .................... | .................... | .................... |
| A | 1296 hCoV-19/France/HDF-2405/2020\|EPI_ISL_418228\|2020-0 | .................... | .................... | .................... | .................... | .................... | .................... |
| A | 1297 hCoV-19/France/IDF-2410/2020\|EPI_ISL_418229\|2020-0 | .................... | .................... | .................... | .................... | .................... | .................... |
| A | 1298 hCoV-19/France/IDF-2420/2020\|EPI_ISL_418230\|2020-0 | .................... | .................... | .................... | .................... | .................... | .................... |
| A | 1299 hCoV-19/France/HDF-2496/2020\|EPI_ISL_418231\|2020-0 | .................... | .................... | .................... | .................... | .................... | .................... |
| A | 1300 hCoV-19/France/IDF-2532/2020\|EPI_ISL_418232\|2020-0 | .................... | .................... | .................... | .................... | .................... | .................... |
| A | 1301 hCoV-19/France/IDF-2533/2020\|EPI_ISL_418233\|2020-0 | .................... | .................... | .................... | .................... | .................... | .................... |
| A | 1302 hCoV-19/France/IDF-2534/2020\|EPI_ISL_418234\|2020-0 | .................... | .................... | .................... | .................... | .................... | .................... |
| A | 1303 hCoV-19/France/IDF-2561/2020\|EPI_ISL_418235\|2020-0 | .................... | .................... | .................... | .................... | .................... | .................... |
| A | 1304 hCoV-19/France/HDF-2586/2020\|EPI_ISL_418236\|2020-0 | .................... | .................... | .................... | .................... | .................... | .................... |
| A | 1305 hCoV-19/France/HDF-2595/2020\|EPI_ISL_418237\|2020-0 | .................... | .................... | .................... | .................... | .................... | .................... |
| A | 1306 hCoV-19/France/HDF-2597/2020\|EPI_ISL_418238\|2020-0 | .................... | .................... | .................... | .................... | .................... | .................... |
| A | 1307 hCoV-19/France/HDF-2601/2020\|EPI_ISL_418239\|2020-0 | .................... | .................... | .................... | .................... | .................... | .................... |
| A | 1308 hCoV-19/France/IDF-2684/2020\|EPI_ISL_418240\|2020-0 | .................... | .................... | .................... | .................... | .................... | .................... |
| A | 1309 hCoV-19/Algeria/G0638_2264/2020\|EPI_ISL_418241\|202 | .................... | .................... | .................... | .................... | .................... | .................... |
| A | 1310 hCoV-19/Algeria/G0640_2265/2020\|EPI_ISL_418242\|202 | .................... | .................... | .................... | .................... | .................... | .................... |
| A | 1311 hCoV-19/Spain/AN-ISCIII-201272/2020\|EPI_ISL_418243 | .................... | .................... | .................... | .................... | .................... | .................... |
| A | 1312 hCoV-19/Spain/AN-ISCIII-201373/2020\|EPI_ISL_418244 | .................... | .................... | .................... | .................... | .................... | .................... |
| A | 1313 hCoV-19/Spain/CM-ISCIII-201328/2020\|EPI_ISL_418245 | .................... | .................... | .................... | .................... | .................... | .................... |
| A | 1314 hCoV-19/Spain/CM-ISCIII-201329/2020\|EPI_ISL_418246 | .................... | .................... | .................... | .................... | .................... | .................... |
| A | 1315 hCoV-19/Spain/CL-ISCIII-201061/2020\|EPI_ISL_418247 | .................... | .................... | .................... | .................... | .................... | .................... |
| A | 1316 hCoV-19/Spain/CL-ISCIII-201323/2020\|EPI_ISL_418248 | .................... | .................... | .................... | .................... | .................... | .................... |
| A | 1317 hCoV-19/Spain/CL-ISCIII-201372/2020\|EPI_ISL_418249 | .................... | .................... | .................... | .................... | .................... | .................... |
| A | 1318 hCoV-19/Spain/CT-ISCIII-201396/2020\|EPI_ISL_418250 | .................... | .................... | .................... | .................... | .................... | .................... |
| A | 1319 hCoV-19/Spain/MD-ISCIII-201105/2020\|EPI_ISL_418251 | .................... | .................... | .................... | .................... | .................... | .................... |
| A | 1320 hCoV-19/Spain/MD-ISCIII-201449/2020\|EPI_ISL_418252 | .................... | .................... | .................... | .................... | .................... | .................... |
| A | 1321 hCoV-19/Spain/PV-ISCIII-201382/2020\|EPI_ISL_418253 | .................... | .................... | .................... | .................... | .................... | .................... |
| A | 1322 hCoV-19/Italy/ABR-IZSGC-TE4925/2020\|EPI_ISL_418255 | .................... | .................... | .................... | .................... | .................... | .................... |
| A | 1323 hCoV-19/Italy/ABR-IZSGC-TE4880/2020\|EPI_ISL_418256 | .................... | .................... | .................... | .................... | .................... | .................... |
| A | 1324 hCoV-19/Italy/ABR-IZSGC-TE5056/2020\|EPI_ISL_418257 | .................... | .................... | .................... | .................... | .................... | .................... |
| A | 1325 hCoV-19/Italy/ABR-IZSGC-TE4953/2020\|EPI_ISL_418258 | .................... | .................... | .................... | .................... | .................... | .................... |
| A | 1327 hCoV-19/Italy/ABR-IZSGC-TE4836/2020\|EPI_ISL_418260 | .................... | .................... | .................... | .................... | .................... | .................... |
| A | 1328 hCoV-19/Colombia/DC-INS-78390/2020\|EPI_ISL_418262\| | .................... | .................... | .................... | .................... | .................... | .................... |
| A | 1329 hCoV-19/Greece/10/2020\|EPI_ISL_418263\|2020-03-18 | .................... | .................... | .................... | .................... | .................... | .................... |

A 1330 hCoV-19/Greece/12/2020|EPI_ISL_418264|2020-03-18 .................... .................... .................... .................... .................... ....................
A 1331 hCoV-19/Greece/16/2020|EPI_ISL_418265|2020-03-18 .................... .................... .................... .................... .................... ....................
A 1332 hCoV-19/Vietnam/19-02S/2020|EPI_ISL_418267|2020-01 .................... .................... .................... .................... .................... ....................
A 1333 hCoV-19/Vietnam/19-01S/2020|EPI_ISL_418269|2020-01 .................... .................... .................... .................... .................... ....................
A 1334 hCoV-19/Belgium/JL-03044/2020|EPI_ISL_418270|2020- .................... .................... .................... .................... .................... ....................
A 1338 hCoV-19/England/SHEF-BFDCD/2020|EPI_ISL_418291|202 .................... .................... .................... .................... .................... ....................
A 1339 hCoV-19/England/SHEF-BFDDC/2020|EPI_ISL_418292|202 .................... .................... .................... .................... .................... ....................
A 1340 hCoV-19/England/SHEF-BFDEB/2020|EPI_ISL_418293|202 .................... .................... .................... .................... .................... ....................
A 1341 hCoV-19/England/SHEF-BFDFA/2020|EPI_ISL_418294|202 .................... .................... .................... .................... .................... ....................
A 1342 hCoV-19/England/SHEF-BFE06/2020|EPI_ISL_418295|202 .................... .................... .................... .................... .................... ....................
A 1343 hCoV-19/England/SHEF-BFE15/2020|EPI_ISL_418296|202 .................... .................... .................... .................... .................... ....................
A 1344 hCoV-19/England/SHEF-BFE24/2020|EPI_ISL_418297|202 .................... .................... .................... .................... .................... ....................
A 1345 hCoV-19/England/SHEF-BFE33/2020|EPI_ISL_418298|202 .................... .................... .................... .................... .................... ....................
A 1346 hCoV-19/England/SHEF-BFE42/2020|EPI_ISL_418299|202 .................... .................... .................... .................... .................... ....................
A 1347 hCoV-19/Germany/NW-HHU-33/2020|EPI_ISL_419550|2020 .................... .................... .................... .................... .................... ....................
A 1348 hCoV-19/Germany/NW-HHU-34/2020|EPI_ISL_419551|2020 .................... .................... .................... .................... .................... ....................
A 1349 hCoV-19/Germany/NW-HHU-35/2020|EPI_ISL_419552|2020 .................... .................... .................... .................... .................... ....................
A 1350 hCoV-19/USA/RI_0520/2020|EPI_ISL_419553|2020-02-28 .................... .................... .................... .................... .................... ....................
A 1351 hCoV-19/USA/CA-CDC-2602/2020|EPI_ISL_419554|2020-0 .................... .................... .................... .................... .................... ....................
A 1352 hCoV-19/USA/WA_5030/2020|EPI_ISL_419555|2020-02-27 .................... .................... .................... .................... .................... ....................
A 1353 hCoV-19/USA/GA_2741/2020|EPI_ISL_419556|2020-02-29 .................... .................... .................... .................... .................... ....................
A 1354 hCoV-19/USA/GA_2742/2020|EPI_ISL_419557|2020-02-29 .................... .................... .................... .................... .................... ....................
A 1355 hCoV-19/USA/OR_2656/2020|EPI_ISL_419558|2020-02-27 .................... .................... .................... .................... .................... ....................
A 1356 hCoV-19/USA/FL_5125/2020|EPI_ISL_419559|2020-02-28 .................... .................... .................... .................... .................... ....................
A 1357 hCoV-19/USA/FL_5091/2020|EPI_ISL_419560|2020-02-28 .................... .................... .................... .................... .................... ....................
A 1358 hCoV-19/Luxembourg/LNS0000001/2020|EPI_ISL_419562| .................... .................... .................... .................... .................... ....................
A 1359 hCoV-19/Luxembourg/LNS0156959/2020|EPI_ISL_419563| .................... .................... .................... .................... .................... ....................
A 1360 hCoV-19/Luxembourg/LNS0366116/2020|EPI_ISL_419564| .................... .................... .................... .................... .................... ....................
A 1361 hCoV-19/Luxembourg/LNS0641910/2020|EPI_ISL_419566| .................... .................... .................... .................... .................... ....................
A 1362 hCoV-19/Luxembourg/LNS0756270/2020|EPI_ISL_419568| .................... .................... .................... .................... .................... ....................
A 1363 hCoV-19/Luxembourg/LNS0945359/2020|EPI_ISL_419569| .................... .................... .................... .................... .................... ....................
A 1364 hCoV-19/Luxembourg/LNS1234709/2020|EPI_ISL_419570| .................... .................... .................... .................... .................... ....................
A 1365 hCoV-19/Luxembourg/LNS1874423/2020|EPI_ISL_419573| .................... .................... .................... .................... .................... ....................
A 1366 hCoV-19/Luxembourg/LNS2614631/2020|EPI_ISL_419578| .................... .................... .................... .................... .................... ....................
A 1367 hCoV-19/Luxembourg/LNS2886370/2020|EPI_ISL_419579| .................... .................... .................... .................... .................... ....................
A 1368 hCoV-19/Luxembourg/LNS2907333/2020|EPI_ISL_419580| .................... .................... .................... .................... .................... ....................
A 1369 hCoV-19/Luxembourg/LNS3156434/2020|EPI_ISL_419582| .................... .................... .................... .................... .................... ....................
A 1370 hCoV-19/Luxembourg/LNS3588186/2020|EPI_ISL_419583| .................... .................... .................... .................... .................... ....................
A 1371 hCoV-19/Luxembourg/LNS3711853/2020|EPI_ISL_419584| .................... .................... .................... .................... .................... ....................
A 1372 hCoV-19/Luxembourg/LNS3879580/2020|EPI_ISL_419585| .................... .................... .................... .................... .................... ....................
A 1373 hCoV-19/Luxembourg/LNS4134806/2020|EPI_ISL_419586| .................... .................... .................... .................... .................... ....................
A 1374 hCoV-19/Luxembourg/LNS4691488/2020|EPI_ISL_419587| .................... .................... .................... .................... .................... ....................
A 1375 hCoV-19/Luxembourg/LNS4836560/2020|EPI_ISL_419588| .................... .................... .................... .................... .................... ....................
A 1376 hCoV-19/Luxembourg/LNS4845603/2020|EPI_ISL_419589| .................... ...................A .................... .................... .................... .................... 1
A 1377 hCoV-19/Luxembourg/LNS5032431/2020|EPI_ISL_419590| .................... .................... .................... .................... .................... ....................
A 1378 hCoV-19/Luxembourg/LNS5228153/2020|EPI_ISL_419591| .................... .................... .................... .................... .................... ....................
A 1379 hCoV-19/Luxembourg/LNS5440244/2020|EPI_ISL_419592| .................... .................... .................... .................... .................... ....................
A 1380 hCoV-19/Luxembourg/LNS6603085/2020|EPI_ISL_419593| .................... .................... .................... .................... .................... ....................
A 1381 hCoV-19/Luxembourg/LNS7537751/2020|EPI_ISL_419594| .................... .................... .................... .................... .................... ....................
A 1382 hCoV-19/Luxembourg/LNS7753431/2020|EPI_ISL_419595| .................... .................... .................... .................... .................... ....................
A 1383 hCoV-19/Luxembourg/LNS7866283/2020|EPI_ISL_419596| .................... .................... .................... .................... .................... ....................
A 1384 hCoV-19/Luxembourg/LNS7928872/2020|EPI_ISL_419597| .................... .................... .................... .................... .................... ....................
A 1385 hCoV-19/Luxembourg/LNS8188502/2020|EPI_ISL_419598| .................... .................... .................... .................... .................... ....................
A 1386 hCoV-19/Luxembourg/LNS8639502/2020|EPI_ISL_419599| .................... .................... .................... .................... .................... ....................
A 1387 hCoV-19/Australia/WA02/2020|EPI_ISL_420532|2020-03 .................... .................... .................... .................... .................... ....................
A 1388 hCoV-19/Australia/WA03/2020|EPI_ISL_420533|2020-03 .................... .................... .................... .................... .................... ....................
A 1389 hCoV-19/Australia/WA06/2020|EPI_ISL_420534|2020-03 .................... .................... .................... .................... .................... ....................
A 1390 hCoV-19/Australia/WA04/2020|EPI_ISL_420536|2020-03 .................... .................... .................... .................... .................... ....................
A 1391 hCoV-19/Australia/WA07/2020|EPI_ISL_420537|2020-03 .................... .................... .................... .................... .................... ....................
A 1392 hCoV-19/Australia/WA09/2020|EPI_ISL_420538|2020-03 .................... .................... .................... .................... .................... ....................
A 1393 hCoV-19/Australia/WA10/2020|EPI_ISL_420539|2020-03 .................... .................... .................... .................... .................... ....................
A 1394 hCoV-19/Estonia/ChVir2148/2020|EPI_ISL_420540|2020 .................... .................... .................... .................... .................... ....................
A 1395 hCoV-19/Slovenia/808/2020|EPI_ISL_420541|2020-03-0 .................... .................... .................... .................... .................... ....................
A 1396 hCoV-19/India/UN-763/2020|EPI_ISL_420543|2020-03-0 .................... .................... .................... .................... .................... ....................
A 1397 hCoV-19/India/UN-2020763/2020|EPI_ISL_420544|2020 .................... .................... .................... .................... .................... ....................
A 1398 hCoV-19/India/UN-770/2020|EPI_ISL_420545|2020-03-0 .................... .................... .................... .................... .................... ....................
A 1399 hCoV-19/India/UN-2020770/2020|EPI_ISL_420546|2020 .................... .................... .................... .................... .................... ....................
A 1400 hCoV-19/India/UN-772/2020|EPI_ISL_420547|2020-03-0 .................... .................... .................... .................... .................... ....................
A 1401 hCoV-19/India/UN-2020772/2020|EPI_ISL_420548|2020 .................... .................... .................... .................... .................... ....................
A 1402 hCoV-19/India/UN-773/2020|EPI_ISL_420549|2020-03-0 .................... .................... .................... .................... .................... ....................
A 1403 hCoV-19/India/UN-2020773/2020|EPI_ISL_420550|2020 .................... .................... .................... .................... .................... ....................
A 1404 hCoV-19/India/UN-777/2020|EPI_ISL_420551|2020-03-0 .................... .................... .................... .................... .................... ....................
A 1405 hCoV-19/India/UN-2020777/2020|EPI_ISL_420552|2020 .................... .................... .................... .................... .................... ....................
A 1406 hCoV-19/India/UN-781/2020|EPI_ISL_420553|2020-03-0 .................... .................... .................... .................... .................... ....................
A 1407 hCoV-19/India/UN-2020781/2020|EPI_ISL_420554|2020 .................... .................... .................... .................... .................... ....................
A 1408 hCoV-19/India/UN-c32/2020|EPI_ISL_420555|2020-03-0 .................... .................... .................... .................... .................... ....................
A 1409 hCoV-19/India/UN-2020c32/2020|EPI_ISL_420556|2020 .................... .................... .................... .................... .................... ....................
A 1410 hCoV-19/Italy/ABR-IZSGC-TE5166/2020|EPI_ISL_420563 .................... .................... .................... .................... .................... ....................

| | | | | | | | | |
|---|---|---|---|---|---|---|---|---|
| A | 1411 hCoV-19/Italy/ABR-IZSGC-TE5472/2020\|EPI_ISL_420564 | ···················· | ···················· | ···················· | ···················· | ···················· | ···················· | |
| A | 1412 hCoV-19/Italy/TE-IZSGC-TE5476/2020\|EPI_ISL_420565\| | ···················· | ···················· | ···················· | ···················· | ···················· | ···················· | |
| A | 1413 hCoV-19/Italy/ABR-IZSGC-TE5512/2020\|EPI_ISL_420566 | ···················· | ···················· | ···················· | ······N············· | ···················· | ···················· | |
| A | 1414 hCoV-19/Italy/ABR-IZSGC-TE5780/2020\|EPI_ISL_420567 | ···················· | ···················· | ···················· | ···················· | ···················· | ···················· | |
| A | 1415 hCoV-19/Italy/ABR-IZSGC-6193/2020\|EPI_ISL_420568\|2 | ···················· | ···················· | ···················· | ···················· | ···················· | ···················· | |
| A | 1416 hCoV-19/Italy/ABR-IZSGC-TE6195/2020\|EPI_ISL_420569 | ···················· | ···················· | ···················· | ···················· | ···················· | ···················· | |
| A | 1417 hCoV-19/USA/NY-NYUMC55/2020\|EPI_ISL_420570\|2020-03 | ···················· | ···················· | ···················· | ···················· | ···················· | ···················· | |
| A | 1418 hCoV-19/USA/NY-NYUMC56/2020\|EPI_ISL_420571\|2020-03 | ···················· | ···················· | ···················· | ···················· | ···················· | ···················· | |
| A | 1419 hCoV-19/USA/NY-NYUMC57/2020\|EPI_ISL_420572\|2020-03 | ···················· | ···················· | ···················· | ···················· | ···················· | ···················· | |
| A | 1420 hCoV-19/USA/NY-NYUMC58/2020\|EPI_ISL_420573\|2020-03 | ···················· | ···················· | ···················· | ···················· | ···················· | ···················· | |
| A | 1421 hCoV-19/USA/NY-NYUMC60/2020\|EPI_ISL_420575\|2020-03 | ···················· | ···················· | ···················· | ···················· | ···················· | ···················· | |
| A | 1422 hCoV-19/USA/NY-NYUMC61/2020\|EPI_ISL_420576\|2020-03 | ···················· | ···················· | ···················· | ···················· | ···················· | ···················· | |
| A | 1423 hCoV-19/USA/NY-NYUMC62/2020\|EPI_ISL_420577\|2020-03 | ···················· | ···················· | ···················· | ···················· | ···················· | ···················· | |
| A | 1424 hCoV-19/USA/NY-NYUMC63/2020\|EPI_ISL_420578\|2020-03 | ···················· | ···················· | ···················· | ···················· | ···················· | ···················· | |
| A | 1425 hCoV-19/USA/NY-NYUMC64/2020\|EPI_ISL_420579\|2020-03 | ···················· | ···················· | ···················· | ···················· | ···················· | ···················· | |
| A | 1426 hCoV-19/USA/NY-NYUMC65/2020\|EPI_ISL_420580\|2020-03 | ···················· | ···················· | ···················· | ···················· | ···················· | ···················· | |
| A | 1427 hCoV-19/USA/NY-NYUMC66/2020\|EPI_ISL_420581\|2020-03 | ···················· | ···················· | ···················· | ···················· | ···················· | ···················· | |
| A | 1428 hCoV-19/USA/NY-NYUMC67/2020\|EPI_ISL_420582\|2020-03 | ···················· | ···················· | ···················· | ···················· | ···················· | ···················· | |
| A | 1429 hCoV-19/Italy/ABR-IZSGC-TE6222/2020\|EPI_ISL_420583 | ···················· | ···················· | ···················· | ···················· | ···················· | ···················· | |
| A | 1430 hCoV-19/USA/NY-NYUMC69/2020\|EPI_ISL_420585\|2020-03 | ···················· | ···················· | ···················· | ···················· | ···················· | ···················· | |
| A | 1431 hCoV-19/USA/NY-NYUMC70/2020\|EPI_ISL_420586\|2020-03 | ···················· | ···················· | ···················· | ···················· | ···················· | ···················· | |
| A | 1432 hCoV-19/USA/NY-NYUMC71/2020\|EPI_ISL_420587\|2020-03 | ···················· | ···················· | ···················· | ···················· | ···················· | ···················· | |
| A | 1433 hCoV-19/USA/NY-NYUMC72/2020\|EPI_ISL_420588\|2020-03 | ···················· | ···················· | ···················· | ···················· | ···················· | ···················· | |
| A | 1434 hCoV-19/USA/NY-NYUMC73/2020\|EPI_ISL_420589\|2020-03 | ···················· | ···················· | ···················· | ···················· | ···················· | ···················· | |
| A | 1435 hCoV-19/USA/NY-NYUMC74/2020\|EPI_ISL_420590\|2020-03 | ···················· | ···················· | ···················· | ···················· | ···················· | ···················· | |
| A | 1436 hCoV-19/USA/NY-NYUMC75/2020\|EPI_ISL_420591\|2020-03 | ···················· | ···················· | ···················· | ···················· | ···················· | ···················· | |
| A | 1437 hCoV-19/England/20139008304/2020\|EPI_ISL_421863\|20 | ···················· | ···················· | ···················· | ···················· | ···················· | ···················· | |
| A | 1438 hCoV-19/England/20139008404/2020\|EPI_ISL_421864\|20 | ···················· | ···················· | ···················· | ···················· | ···················· | ···················· | |
| A | 1439 hCoV-19/England/20139008704/2020\|EPI_ISL_421865\|20 | ···················· | ···················· | ···················· | ···················· | ···················· | ···················· | |
| A | 1440 hCoV-19/England/20139009004/2020\|EPI_ISL_421866\|20 | ···················· | ···················· | ···················· | ···················· | ···················· | ···················· | |
| A | 1441 hCoV-19/England/20139009204/2020\|EPI_ISL_421867\|20 | ···················· | ···················· | ···················· | ···················· | ···················· | ···················· | |
| A | 1442 hCoV-19/England/20139009504/2020\|EPI_ISL_421868\|20 | ···················· | ···················· | ···················· | ···················· | ···················· | ···················· | |
| A | 1445 hCoV-19/England/20139010604/2020\|EPI_ISL_421873\|20 | ···················· | ···················· | ···················· | ···················· | ···················· | ···················· | |
| A | 1448 hCoV-19/England/20139020204/2020\|EPI_ISL_421879\|20 | ···················· | ···················· | ···················· | ···················· | ···················· | ···················· | |
| A | 1449 hCoV-19/England/20139021104/2020\|EPI_ISL_421880\|20 | ···················· | ···················· | ···················· | ···················· | ···················· | ···················· | |
| A | 1450 hCoV-19/England/20139021604/2020\|EPI_ISL_421882\|20 | ···················· | ···················· | ···················· | ···················· | ···················· | ···················· | |
| A | 1451 hCoV-19/England/20139022004/2020\|EPI_ISL_421883\|20 | ···················· | ···················· | ···················· | ···················· | ···················· | ···················· | |
| A | 1452 hCoV-19/England/20139022204/2020\|EPI_ISL_421884\|20 | ···················· | ···················· | ···················· | ···················· | ···················· | ···················· | |
| A | 1453 hCoV-19/England/20139022504/2020\|EPI_ISL_421885\|20 | ···················· | ···················· | ···················· | ···················· | ···················· | ···N················ | |
| A | 1454 hCoV-19/England/20139022704/2020\|EPI_ISL_421886\|20 | ···················· | ···················· | ···················· | ···················· | NNN················· | ···················· | 1 |
| A | 1455 hCoV-19/England/20139023504/2020\|EPI_ISL_421887\|20 | ···················· | ···················· | ···················· | ···················· | ···················· | ···················· | |
| A | 1457 hCoV-19/England/20139023804/2020\|EPI_ISL_421889\|20 | ···················· | ···················· | ···················· | ···················· | ···················· | ···················· | |
| A | 1458 hCoV-19/England/20139024004/2020\|EPI_ISL_421890\|20 | ···················· | ···················· | ···················· | ···················· | ···················· | ···················· | |
| A | 1462 hCoV-19/England/20139024804/2020\|EPI_ISL_421894\|20 | ···················· | ···················· | ···················· | ···················· | ···················· | ···················· | |
| A | 1464 hCoV-19/England/20139026904/2020\|EPI_ISL_421899\|20 | ···················· | ···················· | ···················· | ···················· | ···················· | ···················· | |
| A | 1465 hCoV-19/USA/WA-UW-1420/2020\|EPI_ISL_423000\|2020-03 | ···················· | ···················· | ···················· | ···················· | ···················· | ···················· | |
| A | 1466 hCoV-19/USA/WA-UW-1567/2020\|EPI_ISL_423001\|2020-03 | ···················· | ···················· | ···················· | ···················· | ···················· | ···················· | |
| A | 1467 hCoV-19/USA/WA-UW-1593/2020\|EPI_ISL_423002\|2020-03 | ···················· | ···················· | ···················· | ···················· | ···················· | ···················· | |
| A | 1468 hCoV-19/USA/WA-UW-1583/2020\|EPI_ISL_423003\|2020-03 | ···················· | ···················· | ···················· | ···················· | ···················· | ···················· | |
| A | 1469 hCoV-19/USA/WA-UW-1581/2020\|EPI_ISL_423004\|2020-03 | ···················· | ···················· | ···················· | ···················· | ···················· | ···················· | |
| A | 1470 hCoV-19/USA/WA-UW-1603/2020\|EPI_ISL_423005\|2020-03 | ···················· | ···················· | ···················· | ···················· | ···················· | ···················· | |
| A | 1471 hCoV-19/USA/WA-UW-1611/2020\|EPI_ISL_423006\|2020-03 | ···················· | ···················· | ···················· | ···················· | ···················· | ···················· | |
| A | 1472 hCoV-19/USA/WA-UW-1879/2020\|EPI_ISL_424300\|2020-03 | ···················· | ···················· | ···················· | ···················· | ···················· | ···················· | |
| A | 1473 hCoV-19/USA/WA-UW-1948/2020\|EPI_ISL_424301\|2020-03 | ···················· | ···················· | ···················· | ···················· | ···················· | ···················· | |
| A | 1474 hCoV-19/USA/WA-UW-1890/2020\|EPI_ISL_424316\|2020-03 | ···················· | ···················· | ···················· | ···················· | ···················· | ···················· | |
| A | 1475 hCoV-19/USA/WA-UW-1928/2020\|EPI_ISL_424317\|2020-03 | ···················· | ···················· | ···················· | ···················· | ···················· | ···················· | |
| A | 1476 hCoV-19/USA/ID-UW-1938/2020\|EPI_ISL_424318\|2020-03 | ···················· | ···················· | ···················· | ···················· | ···················· | ···················· | |
| A | 1477 hCoV-19/USA/ID-UW-1935/2020\|EPI_ISL_424319\|2020-03 | ···················· | ···················· | ···················· | ···················· | ···················· | ···················· | |
| A | 1478 hCoV-19/USA/ID-UW-1925/2020\|EPI_ISL_424321\|2020-03 | ···················· | ···················· | ···················· | ···················· | ···················· | ···················· | |
| A | 1479 hCoV-19/USA/ID-UW-1934/2020\|EPI_ISL_424322\|2020-03 | ···················· | ···················· | ···················· | ···················· | ···················· | ···················· | |
| A | 1480 hCoV-19/USA/WA-UW-1944/2020\|EPI_ISL_424323\|2020-03 | ···················· | ···················· | ···················· | ···················· | ···················· | ···················· | |
| A | 1481 hCoV-19/USA/WA-UW-1965/2020\|EPI_ISL_424324\|2020-03 | ···················· | ···················· | ···················· | ···················· | ···················· | ···················· | |
| A | 1482 hCoV-19/USA/WA-UW-1954/2020\|EPI_ISL_424325\|2020-03 | ···················· | ···················· | ···················· | ···················· | ···················· | ···················· | |
| A | 1483 hCoV-19/USA/WA-UW-1968/2020\|EPI_ISL_424326\|2020-03 | ···················· | ···················· | ···················· | ···················· | ···················· | ···················· | |
| A | 1484 hCoV-19/USA/WA-UW-1984/2020\|EPI_ISL_424329\|2020-03 | ···················· | ···················· | ···················· | ···················· | ···················· | ···················· | |
| A | 1485 hCoV-19/USA/WA-UW-1865/2020\|EPI_ISL_424330\|2020-03 | ···················· | ···················· | ···················· | ···················· | ···················· | ···················· | |
| A | 1486 hCoV-19/USA/WA-UW-1907/2020\|EPI_ISL_424331\|2020-03 | ···················· | ···················· | ···················· | ···················· | ···················· | ···················· | |
| A | 1487 hCoV-19/USA/WA-UW-1917/2020\|EPI_ISL_424332\|2020-03 | ···················· | ···················· | ···················· | ···················· | ···················· | ···················· | |
| A | 1488 hCoV-19/USA/WA-UW-2006/2020\|EPI_ISL_424333\|2020-03 | ···················· | ···················· | ···················· | ···················· | ···················· | ···················· | |
| A | 1489 hCoV-19/USA/WA-UW-1994/2020\|EPI_ISL_424334\|2020-03 | ···················· | ···················· | ···················· | ···················· | ···················· | ···················· | |
| A | 1490 hCoV-19/USA/WA-UW-1998/2020\|EPI_ISL_424335\|2020-03 | ···················· | ···················· | ···················· | ···················· | ···················· | ···················· | |
| A | 1491 hCoV-19/USA/WA-UW-1988/2020\|EPI_ISL_424336\|2020-03 | ···················· | ···················· | ···················· | ···················· | ···················· | ···················· | |
| A | 1492 hCoV-19/USA/WA-UW-1977/2020\|EPI_ISL_424337\|2020-03 | ···················· | ···················· | ···················· | ···················· | ···················· | ···················· | |
| A | 1493 hCoV-19/England/NOTT-10EA21/2020\|EPI_ISL_425627\|20 | ···················· | ···················· | ···················· | ···················· | ···················· | ···················· | |
| A | 1494 hCoV-19/England/NOTT-10EA30/2020\|EPI_ISL_425628\|20 | ···················· | ···················· | ···················· | ···················· | ···················· | ···················· | |
| A | 1495 hCoV-19/England/NOTT-10EA5E/2020\|EPI_ISL_425630\|20 | ···················· | ···················· | ···················· | ···················· | ···················· | ···················· | |
| A | 1496 hCoV-19/England/NOTT-10EA6D/2020\|EPI_ISL_425631\|20 | ···················· | ···················· | ···················· | ···················· | ···················· | ···················· | |
| A | 1497 hCoV-19/England/NOTT-10EA7C/2020\|EPI_ISL_425632\|20 | ···················· | ···················· | ···················· | ···················· | ···················· | ···················· | |

| | | | | | | | |
|---|---|---|---|---|---|---|---|
| A | 1498 hCoV-19/Scotland/CVR100/2020\|EPI_ISL_425647\|2020-0 | ···················· | ···················· | ····················· | ···················· | ···················· | ···················· |
| A | 1499 hCoV-19/Scotland/CVR101/2020\|EPI_ISL_425648\|2020-0 | ···················· | ···················· | ····················· | ···················· | ···················· | ···················· |
| A | 1500 hCoV-19/Scotland/CVR103/2020\|EPI_ISL_425650\|2020-0 | ···················· | ···················· | ····················· | ···················· | ···················· | ···················· |
| A | 1501 hCoV-19/Scotland/CVR104/2020\|EPI_ISL_425651\|2020-0 | ···················· | ···················· | ····················· | ···················· | ···················· | ···················· |
| A | 1502 hCoV-19/Scotland/CVR106/2020\|EPI_ISL_425652\|2020-0 | ···················· | ···················· | ····················· | ···················· | ···················· | ···················· |
| A | 1503 hCoV-19/Scotland/CVR107/2020\|EPI_ISL_425653\|2020-0 | ···················· | ···················· | ····················· | ···················· | ···················· | ···················· |
| A | 1504 hCoV-19/Scotland/CVR108/2020\|EPI_ISL_425654\|2020-0 | ···················· | ···················· | ····················· | ···················· | ···················· | ···················· |
| A | 1505 hCoV-19/Scotland/CVR110/2020\|EPI_ISL_425656\|2020-0 | ···················· | ···················· | ····················· | ···················· | ···················· | ···················· |
| A | 1506 hCoV-19/Scotland/CVR112/2020\|EPI_ISL_425657\|2020-0 | ···················· | ···················· | ····················· | ···················· | ···················· | ···················· |
| A | 1507 hCoV-19/Scotland/CVR113/2020\|EPI_ISL_425658\|2020-0 | ···················· | ···················· | ····················· | ···················· | ···················· | ···················· |
| A | 1508 hCoV-19/Scotland/CVR114/2020\|EPI_ISL_425659\|2020-0 | ···················· | ···················· | ····················· | ···················· | ···················· | ···················· |
| A | 1509 hCoV-19/Scotland/CVR115/2020\|EPI_ISL_425660\|2020-0 | ···················· | ···················· | ····················· | ···················· | ···················· | ···················· |
| A | 1510 hCoV-19/Scotland/CVR118/2020\|EPI_ISL_425662\|2020-0 | ···················· | ···················· | ····················· | ···················· | ···················· | ···················· |
| A | 1511 hCoV-19/Scotland/CVR12/2020\|EPI_ISL_425663\|2020-03 | ···················· | ···················· | ····················· | ···················· | ···················· | ···················· |
| A | 1512 hCoV-19/Scotland/CVR125/2020\|EPI_ISL_425668\|2020-0 | ···················· | ···················· | ····················· | ···················· | ···················· | ···················· |
| A | 1514 hCoV-19/Australia/VIC661/2020\|EPI_ISL_426957\|2020- | ···················· | ···················· | ····················· | ···················· | ···················· | ···················· |
| A | 1515 hCoV-19/Australia/VIC662/2020\|EPI_ISL_426958\|2020- | ···················· | ···················· | ····················· | ···················· | ···················· | ···················· |
| A | 1517 hCoV-19/Australia/VIC664/2020\|EPI_ISL_426960\|2020- | ···················· | ···················· | ····················· | ···················· | ···················· | ···················· |
| A | 1518 hCoV-19/Australia/VIC665/2020\|EPI_ISL_426961\|2020- | ···················· | ···················· | ····················· | ···················· | ···················· | ···················· |
| A | 1520 hCoV-19/Australia/VIC667/2020\|EPI_ISL_426963\|2020- | ···················· | ···················· | ····················· | ···················· | ···················· | ···················· |
| A | 1521 hCoV-19/Australia/VIC668/2020\|EPI_ISL_426964\|2020- | ···················· | ···················· | ····················· | ···················· | ···················· | ···················· |
| A | 1522 hCoV-19/Australia/VIC669/2020\|EPI_ISL_426965\|2020- | ···················· | ···················· | ····················· | ···················· | ···················· | ···················· |
| A | 1523 hCoV-19/Australia/VIC672/2020\|EPI_ISL_426968\|2020- | ···················· | ···················· | ····················· | ···················· | ···················· | ···················· |
| A | 1525 hCoV-19/Australia/VIC676/2020\|EPI_ISL_426971\|2020- | ···················· | ···················· | ····················· | ···················· | ···················· | ···················· |
| A | 1526 hCoV-19/Australia/VIC677/2020\|EPI_ISL_426972\|2020- | ···················· | ···················· | ····················· | ···················· | ···················· | ···················· |
| A | 1528 hCoV-19/Australia/VIC679/2020\|EPI_ISL_426974\|2020- | ···················· | ···················· | ····················· | ···················· | ···················· | ···················· |
| A | 1529 hCoV-19/Australia/VIC680/2020\|EPI_ISL_426975\|2020- | ···················· | ···················· | ····················· | ···················· | ···················· | ···················· |
| A | 1530 hCoV-19/Australia/VIC681/2020\|EPI_ISL_426976\|2020- | ···················· | ···················· | ····················· | ···················· | ···················· | ···················· |
| A | 1531 hCoV-19/Australia/VIC682/2020\|EPI_ISL_426977\|2020- | ···················· | ···················· | ····················· | ···················· | ···················· | ···················· |
| A | 1535 hCoV-19/Australia/VIC710/2020\|EPI_ISL_426997\|2020- | ···················· | ···················· | ····················· | ···················· | ···················· | ···················· |
| A | 1536 hCoV-19/Australia/VIC712/2020\|EPI_ISL_426999\|2020- | ···················· | ···················· | ····················· | ···················· | ···················· | ···················· |
| A | 1537 hCoV-19/Wuhan/IPBCAMS-WH-05/2020\|EPI_ISL_403928\|20 | ···················· | ···················· | ····················· | ···················· | ···················· | ···················· |
| A | 1538 hCoV-19/Wuhan/IPBCAMS-WH-04/2019\|EPI_ISL_403929\|20 | ···················· | ···················· | ····················· | ···················· | ···················· | ···················· |
| A | 1539 hCoV-19/Guangdong/20SF013/2020\|EPI_ISL_403933\|2020 | ···················· | ···················· | ····················· | ···················· | ···················· | ···················· |
| A | 1540 hCoV-19/Guangdong/20SF014/2020\|EPI_ISL_403934\|2020 | ···················· | ···················· | ····················· | ···················· | ···················· | ···················· |
| A | 1541 hCoV-19/Guangdong/20SF025/2020\|EPI_ISL_403935\|2020 | ···················· | ···················· | ····················· | ···················· | ···················· | ···················· |
| A | 1542 hCoV-19/Guangdong/20SF028/2020\|EPI_ISL_403936\|2020 | ···················· | ···················· | ····················· | ···················· | ···················· | ···················· |
| A | 1543 hCoV-19/Guangdong/20SF040/2020\|EPI_ISL_403937\|2020 | ···················· | ···················· | ····················· | ···················· | ···················· | ···················· |
| A | 1544 hCoV-19/USA/AZ1/2020\|EPI_ISL_406223\|2020-01-22 | ···················· | ···················· | ····················· | ···················· | ···················· | ···················· |
| A | 1545 hCoV-19/Hefei/2/2020\|EPI_ISL_412026\|2020-02-23 | ···················· | ···················· | ····················· | ···················· | ···················· | ···················· |
| A | 1546 hCoV-19/Hong Kong/VM20001061-2/2020\|EPI_ISL_412028 | ···················· | ···················· | ····················· | ···················· | ···················· | ···················· |
| A | 1547 hCoV-19/Hong Kong/VB20024950-2/2020\|EPI_ISL_412029 | ···················· | ···················· | ····················· | ···················· | ···················· | ···················· |
| A | 1548 hCoV-19/Hong Kong/VB20026565-2/2020\|EPI_ISL_412030 | ···················· | ···················· | ····················· | ···················· | ···················· | ···················· |
| A | 1549 hCoV-19/Guangzhou/GZMU0016/2020\|EPI_ISL_414663\|202 | ···················· | ···················· | ····················· | ···················· | ···················· | ···················· |
| A | 1550 hCoV-19/Guangzhou/GZMU0030/2020\|EPI_ISL_414686\|202 | ···················· | ···················· | ····················· | ···················· | ···················· | ···················· |
| A | 1551 hCoV-19/Guangzhou/GZMU0031/2020\|EPI_ISL_414687\|202 | ···················· | ···················· | ····················· | ···················· | ···················· | ···················· |
| A | 1552 hCoV-19/Guangzhou/GZMU0044/2020\|EPI_ISL_414689\|202 | ···················· | ···················· | ····················· | ···················· | ···················· | ···················· |
| A | 1553 hCoV-19/Guangzhou/GZMU0047/2020\|EPI_ISL_414690\|202 | ···················· | ···················· | ····················· | ···················· | ···················· | ···················· |
| A | 1554 hCoV-19/Guangzhou/GZMU0048/2020\|EPI_ISL_414691\|202 | ···················· | ···················· | ····················· | ···················· | ···················· | ···················· |
| A | 1555 hCoV-19/Guangzhou/GZMU0014/2020\|EPI_ISL_414692\|202 | ···················· | ···················· | ····················· | ···················· | ···················· | ···················· |
| A | 1556 hCoV-19/Wales/PHW32/2020\|EPI_ISL_415920\|2020-03-12 | ···················· | ····A··············· | ····················· | ···················· | ···················· | ···················· |
| A | 1557 hCoV-19/USA/WI-GMF-00018/2020\|EPI_ISL_418184\|2020- | ···················· | ···················· | ····················· | ···················· | ···················· | ···················· |
| A | 1558 hCoV-19/USA/WI-GMF-00227/2020\|EPI_ISL_418186\|2020- | ···················· | ···················· | ····················· | ···················· | ···················· | ···················· |
| A | 1559 hCoV-19/USA/WI-GMF-00228/2020\|EPI_ISL_418187\|2020- | ···················· | ···················· | ····················· | ···················· | ···················· | ···················· |
| A | 1560 hCoV-19/USA/WI-GMF-00232/2020\|EPI_ISL_418188\|2020- | ···················· | ···················· | ····················· | ···················· | ···················· | ···················· |
| A | 1561 hCoV-19/USA/NY-NYUMC7/2020\|EPI_ISL_418192\|2020-03- | ···················· | ···················· | ····················· | ···················· | ···················· | ···················· |
| A | 1562 hCoV-19/USA/NY-NYUMC8/2020\|EPI_ISL_418193\|2020-03- | ···················· | ···················· | ····················· | ···················· | ···················· | ···················· |
| A | 1563 hCoV-19/USA/NY-NYUMC11/2020\|EPI_ISL_418196\|2020-03 | ···················· | ···················· | ····················· | ···················· | ···················· | ···················· |
| A | 1564 hCoV-19/USA/NY-NYUMC12/2020\|EPI_ISL_418197\|2020-03 | ···················· | ···················· | ····················· | ···················· | ···················· | ···················· |
| A | 1565 hCoV-19/USA/NY-NYUMC13/2020\|EPI_ISL_418198\|2020-03 | ···················· | ···················· | ····················· | ···················· | ···················· | ···················· |
| A | 1566 hCoV-19/USA/NY-NYUMC14/2020\|EPI_ISL_418199\|2020-03 | ···················· | ···················· | ····················· | ···················· | ···················· | ···················· |
| A | 1567 hCoV-19/USA/NY-NYUMC15/2020\|EPI_ISL_418200\|2020-03 | ···················· | ···················· | ····················· | ···················· | ···················· | ···················· |
| A | 1568 hCoV-19/USA/NY-NYUMC18/2020\|EPI_ISL_418203\|2020-03 | ···················· | ···················· | ····················· | ···················· | ···················· | ···················· |
| A | 1569 hCoV-19/USA/NY-NYUMC19/2020\|EPI_ISL_418204\|2020-03 | ···················· | ···················· | ····················· | ···················· | ···················· | ···················· |
| A | 1570 hCoV-19/Senegal/003/2020\|EPI_ISL_418206\|2020-02-28 | ···················· | ···················· | ····················· | ···················· | ···················· | ···················· |
| A | 1571 hCoV-19/Senegal/016/2020\|EPI_ISL_418207\|2020-03-02 | ···················· | ···················· | ····················· | ···················· | ···················· | ···················· |
| A | 1572 hCoV-19/Senegal/020/2020\|EPI_ISL_418208\|2020-03-04 | ···················· | ···················· | ····················· | ···················· | ···················· | ···················· |
| A | 1573 hCoV-19/Senegal/026/2020\|EPI_ISL_418209\|2020-03-03 | ···················· | ···················· | ····················· | ···················· | ···················· | ···················· |
| A | 1574 hCoV-19/Senegal/073/2020\|EPI_ISL_418210\|2020-03-10 | ···················· | ···················· | ····················· | ···················· | ···················· | ···················· |
| A | 1575 hCoV-19/Senegal/082/2020\|EPI_ISL_418211\|2020-03-11 | ···················· | ···················· | ····················· | ···················· | ···················· | ···················· |
| A | 1576 hCoV-19/Senegal/087/2020\|EPI_ISL_418212\|2020-03-11 | ···················· | ···················· | ····················· | ···················· | ···················· | ···················· |
| A | 1577 hCoV-19/Senegal/094/2020\|EPI_ISL_418213\|2020-03-12 | ···················· | ···················· | ····················· | ···················· | ···················· | ···················· |
| A | 1578 hCoV-19/Senegal/119/2020\|EPI_ISL_418215\|2020-03-12 | ···················· | ···················· | ····················· | ···················· | ···················· | ···················· |
| A | 1579 hCoV-19/Senegal/136/2020\|EPI_ISL_418216\|2020-03-13 | ···················· | ···················· | ····················· | ···················· | ···················· | ···················· |
| A | 1580 hCoV-19/Senegal/139/2020\|EPI_ISL_418217\|2020-03-13 | ···················· | ···················· | ····················· | ···················· | ···················· | ···················· |
| A | 1581 hCoV-19/France/HDF-1465/2020\|EPI_ISL_418218\|2020-0 | ···················· | ···················· | ····················· | ···················· | ···················· | ···················· |
| A | 1582 hCoV-19/France/BRE-1623/2020\|EPI_ISL_418219\|2020-0 | ···················· | ···················· | ····················· | ···················· | ···················· | ···················· |
| A | 1585 hCoV-19/England/SHEF-BFD63/2020\|EPI_ISL_418286\|202 | ···················· | ···················· | ····················· | ···················· | ···················· | ···················· |

| | | | | | | | | |
|---|---|---|---|---|---|---|---|---|
| A | 1586 | hCoV-19/England/SHEF-BFD72/2020\|EPI_ISL_418287\|202 | ···················· | ···················· | ···················· | ···················· | ···················· | ···················· |
| A | 1587 | hCoV-19/England/SHEF-BFD81/2020\|EPI_ISL_418288\|202 | ···················· | ···················· | ···················· | ···················· | ···················· | ···················· |
| A | 1588 | hCoV-19/England/SHEF-BFDAF/2020\|EPI_ISL_418289\|202 | ···················· | ···················· | ···················· | ···················· | ···················· | ···················· |
| A | 1589 | hCoV-19/England/SHEF-BFDBE/2020\|EPI_ISL_418290\|202 | ···················· | ···················· | ···················· | ···················· | ···················· | ···················· |
| A | 1590 | hCoV-19/Wales/PHWC-24D87/2020\|EPI_ISL_419508\|2020- | ···················· | ····A··············· | ···················· | ···················· | ···················· | ···················· |
| A | 1591 | hCoV-19/USA/CT-Yale-013/2020\|EPI_ISL_419517\|2020-0 | ···················· | ···················· | ···················· | ···················· | ···················· | ···················· |
| A | 1592 | hCoV-19/USA/CT-Yale-014/2020\|EPI_ISL_419518\|2020-0 | ···················· | ···················· | ···················· | ···················· | ···················· | ···················· |
| A | 1593 | hCoV-19/USA/CT-Yale-034/2020\|EPI_ISL_419524\|2020-0 | ···················· | ···················· | ···················· | ···················· | ···················· | ···················· |
| A | 1594 | hCoV-19/USA/CT-Yale-036/2020\|EPI_ISL_419525\|2020-0 | ···················· | ···················· | ···················· | ···················· | ···················· | ···················· |
| A | 1595 | hCoV-19/USA/CT-Yale-040/2020\|EPI_ISL_419528\|2020-0 | ···················· | ···················· | ···················· | ···················· | ···················· | ···················· |
| A | 1596 | hCoV-19/Germany/NW-HHU-24/2020\|EPI_ISL_419541\|2020 | ···················· | ···················· | ···················· | ···················· | ···················· | ···················· |
| A | 1597 | hCoV-19/Germany/NW-HHU-25/2020\|EPI_ISL_419542\|2020 | ···················· | ···················· | ···················· | ···················· | ···················· | ···················· |
| A | 1598 | hCoV-19/Germany/NW-HHU-26/2020\|EPI_ISL_419543\|2020 | ···················· | ···················· | ···················· | ···················· | ···················· | ···················· |
| A | 1599 | hCoV-19/Germany/NW-HHU-27/2020\|EPI_ISL_419544\|2020 | ···················· | ···················· | ···················· | ···················· | ···················· | ···················· |
| A | 1600 | hCoV-19/Germany/NW-HHU-28/2020\|EPI_ISL_419545\|2020 | ···················· | ···················· | ···················· | ···················· | ···················· | ···················· |
| A | 1601 | hCoV-19/Germany/NW-HHU-29/2020\|EPI_ISL_419546\|2020 | ···················· | ···················· | ···················· | ···················· | ···················· | ···················· |
| A | 1602 | hCoV-19/Germany/NW-HHU-30/2020\|EPI_ISL_419547\|2020 | ···················· | ···················· | ···················· | ···················· | ···················· | ···················· |
| A | 1603 | hCoV-19/Germany/NW-HHU-31/2020\|EPI_ISL_419548\|2020 | ···················· | ···················· | ···················· | ···················· | ···················· | ···················· |
| A | 1604 | hCoV-19/Germany/NW-HHU-32/2020\|EPI_ISL_419549\|2020 | ···················· | ···················· | ···················· | ···················· | ···················· | ···················· |
| A | 1605 | hCoV-19/England/20132072802/2020\|EPI_ISL_420508\|20 | ···················· | ···················· | ···················· | ···················· | ···················· | ···················· |
| A | 1606 | hCoV-19/England/20136015502/2020\|EPI_ISL_420520\|20 | ···················· | ···················· | ···················· | ···················· | ···················· | ···················· |
| A | 1607 | hCoV-19/England/20138012802/2020\|EPI_ISL_420521\|20 | ···················· | ···················· | ···················· | ···················· | ···················· | ···················· |
| A | 1608 | hCoV-19/England/20138014402/2020\|EPI_ISL_420522\|20 | ···················· | ···················· | ···················· | ···················· | ···················· | ···················· |
| A | 1609 | hCoV-19/England/20139018202/2020\|EPI_ISL_420523\|20 | ···················· | ···················· | ···················· | ···················· | ···················· | ···················· |
| A | 1610 | hCoV-19/England/20139018302/2020\|EPI_ISL_420524\|20 | ···················· | ···················· | ···················· | ···················· | ···················· | ···················· |
| A | 1611 | hCoV-19/England/20139051702/2020\|EPI_ISL_420529\|20 | ···················· | ···················· | ···················· | ···················· | ···················· | ···················· |
| A | 1612 | hCoV-19/Australia/WA01/2020\|EPI_ISL_420531\|2020-03 | ···················· | ···················· | ···················· | ···················· | ···················· | ···················· |
| A | 1613 | hCoV-19/Argentina/C3013/2020\|EPI_ISL_420598\|2020-0 | ···················· | ···········A········ | ···················· | ···················· | ···················· | ···················· |
| A | 1614 | hCoV-19/Argentina/C1374/2020\|EPI_ISL_420599\|2020-0 | ···················· | ···················· | ···················· | ···················· | ···················· | ···················· |
| A | 1615 | hCoV-19/England/201380036/2020\|EPI_ISL_421800\|2020 | ···················· | ···················· | ···················· | ···················· | ···················· | ···················· |
| A | 1616 | hCoV-19/England/201380037/2020\|EPI_ISL_421801\|2020 | ···················· | ···················· | ···················· | ···················· | ···················· | ···················· |
| A | 1617 | hCoV-19/England/201380040/2020\|EPI_ISL_421802\|2020 | ···················· | ···················· | ···················· | ···················· | ···················· | ···················· |
| A | 1618 | hCoV-19/England/201380041/2020\|EPI_ISL_421803\|2020 | ···················· | ···················· | ···················· | ···················· | ···················· | ···················· |
| A | 1619 | hCoV-19/England/201380042/2020\|EPI_ISL_421804\|2020 | ···················· | ···················· | ···················· | ···················· | ··NN················ | ···················· |
| A | 1620 | hCoV-19/England/201380044/2020\|EPI_ISL_421805\|2020 | ···················· | ···················· | ···················· | ···················· | ···················· | ···················· |
| A | 1621 | hCoV-19/England/201380049/2020\|EPI_ISL_421806\|2020 | ···················· | ···················· | ···················· | ···················· | ···················· | ···················· |
| A | 1622 | hCoV-19/England/201380050/2020\|EPI_ISL_421807\|2020 | ···················· | ···················· | ···················· | ···················· | ···················· | ···················· |
| A | 1623 | hCoV-19/England/201380054/2020\|EPI_ISL_421809\|2020 | ···················· | ···················· | ···················· | ···················· | ···················· | ···················· |
| A | 1624 | hCoV-19/England/201380055/2020\|EPI_ISL_421810\|2020 | ···················· | ···················· | ···················· | ···················· | ···················· | ···················· |
| A | 1625 | hCoV-19/England/201380056/2020\|EPI_ISL_421811\|2020 | ···················· | ···················· | ···················· | ···················· | ···················· | ···················· |
| A | 1626 | hCoV-19/England/201380061/2020\|EPI_ISL_421813\|2020 | ···T················ | ···················· | ···················· | ···················· | ···················· | ···················· |
| A | 1627 | hCoV-19/England/201380063/2020\|EPI_ISL_421815\|2020 | ···················· | ···················· | ···················· | ···················· | ···················· | ···················· |
| A | 1628 | hCoV-19/England/201380064/2020\|EPI_ISL_421816\|2020 | ···················· | ···················· | ···················· | ···················· | ···················· | ···················· |
| A | 1630 | hCoV-19/England/201380070/2020\|EPI_ISL_421818\|2020 | ···················· | ···················· | ···················· | ···················· | ···················· | ···················· |
| A | 1631 | hCoV-19/England/201380217/2020\|EPI_ISL_421822\|2020 | ···················· | ···················· | ···················· | ···················· | ···················· | ···················· |
| A | 1632 | hCoV-19/England/201380221/2020\|EPI_ISL_421823\|2020 | ···················· | ···················· | ···················· | ···················· | ···················· | ···················· |
| A | 1633 | hCoV-19/England/201380228/2020\|EPI_ISL_421824\|2020 | ···················· | ···················· | ···················· | ···················· | ···················· | ···················· |
| A | 1634 | hCoV-19/England/201380229/2020\|EPI_ISL_421825\|2020 | ···················· | ···················· | ···················· | ···················· | ···················· | ···················· |
| A | 1635 | hCoV-19/England/201380276/2020\|EPI_ISL_421826\|2020 | ···················· | ···················· | ···················· | ···················· | ···················· | ···················· |
| A | 1637 | hCoV-19/England/201380280/2020\|EPI_ISL_421828\|2020 | ···················· | ···················· | ···················· | ···················· | ···················· | ···················· |
| A | 1641 | hCoV-19/England/20138082004/2020\|EPI_ISL_421834\|20 | ···················· | ···················· | ···················· | ···················· | ···················· | ···················· |
| A | 1642 | hCoV-19/England/20138082704/2020\|EPI_ISL_421836\|20 | ···················· | ···················· | ···················· | ···················· | ···················· | ····N··············· |
| A | 1643 | hCoV-19/England/20138082804/2020\|EPI_ISL_421837\|20 | ···················· | ···················· | ···················· | ···················· | ···················· | ···················· |
| A | 1644 | hCoV-19/England/20138083204/2020\|EPI_ISL_421839\|20 | ···T················ | ···················· | ···················· | ···················· | ···················· | ···················· |
| A | 1646 | hCoV-19/England/20138086904/2020\|EPI_ISL_421842\|20 | ···················· | ···················· | ···················· | ···················· | ···················· | ···················· |
| A | 1647 | hCoV-19/England/20138088004/2020\|EPI_ISL_421845\|20 | ···················· | ···················· | ···················· | ···················· | ···················· | ···················· |
| A | 1648 | hCoV-19/England/20138088204/2020\|EPI_ISL_421847\|20 | ···················· | ···················· | ···················· | ···················· | ···················· | ···················· |
| A | 1649 | hCoV-19/England/20139003004/2020\|EPI_ISL_421848\|20 | ···················· | ···················· | ···················· | ···················· | ···················· | ···················· |
| A | 1650 | hCoV-19/England/20139003404/2020\|EPI_ISL_421850\|20 | ···················· | ···················· | ···················· | ···················· | ···················· | ···················· |
| A | 1651 | hCoV-19/England/20139007204/2020\|EPI_ISL_421859\|20 | ···················· | ···················· | ···················· | ···················· | ···················· | ···················· |
| A | 1652 | hCoV-19/England/20139007604/2020\|EPI_ISL_421861\|20 | ···················· | ···················· | ···················· | ···················· | ···················· | ···················· |
| A | 1653 | hCoV-19/England/20139008204/2020\|EPI_ISL_421862\|20 | ···················· | ···················· | ···················· | ···················· | ···················· | ···················· |
| A | 1655 | hCoV-19/England/20129017104/2020\|EPI_ISL_424003\|20 | ···················· | ···················· | ···················· | ···················· | ···················· | ··N················· |
| A | 1657 | hCoV-19/England/20129018904/2020\|EPI_ISL_424007\|20 | ···················· | ···················· | ···················· | ···················· | ···················· | ···················· |
| A | 1658 | hCoV-19/England/20129020004/2020\|EPI_ISL_424009\|20 | ···················· | ···················· | ···················· | ···················· | ···················· | ···················· |
| A | 1659 | hCoV-19/England/20129020304/2020\|EPI_ISL_424010\|20 | ···················· | ···················· | ···················· | ···················· | ···················· | ···················· |
| A | 1660 | hCoV-19/England/20129020604/2020\|EPI_ISL_424012\|20 | ···················· | ···················· | ···················· | ···················· | ···················· | ···················· |
| A | 1662 | hCoV-19/England/20129025404/2020\|EPI_ISL_424017\|20 | ···················· | ···················· | ···················· | ···················· | ···················· | ···················· |
| A | 1664 | hCoV-19/England/20129032104/2020\|EPI_ISL_424020\|20 | ···················· | ···················· | ···················· | ···················· | ···················· | ···················· |
| A | 1666 | hCoV-19/England/20129033804/2020\|EPI_ISL_424024\|20 | ···················· | ···················· | ···················· | ···················· | ···················· | ···················· |
| A | 1667 | hCoV-19/England/20129033904/2020\|EPI_ISL_424025\|20 | ···················· | ···················· | ···················· | ···················· | ···················· | ···················· |
| A | 1668 | hCoV-19/England/20129035304/2020\|EPI_ISL_424028\|20 | ···················· | ···················· | ···················· | ···················· | ···················· | ···················· |
| A | 1669 | hCoV-19/England/20129035404/2020\|EPI_ISL_424029\|20 | ···················· | ···················· | ···················· | ···················· | ···················· | ···················· |
| A | 1670 | hCoV-19/England/20129035804/2020\|EPI_ISL_424031\|20 | ···················· | ···················· | ···················· | ···················· | ···················· | ···················· |
| A | 1671 | hCoV-19/England/20129036004/2020\|EPI_ISL_424032\|20 | ···················· | ···················· | ···················· | ···················· | ···················· | ···················· |
| A | 1672 | hCoV-19/England/20129036404/2020\|EPI_ISL_424034\|20 | ···················· | ···················· | ···················· | ···················· | ···················· | ···················· |
| A | 1673 | hCoV-19/England/20129036504/2020\|EPI_ISL_424035\|20 | ···················· | ···················· | ···················· | ···················· | ···················· | ···················· |
| A | 1674 | hCoV-19/England/20129037204/2020\|EPI_ISL_424037\|20 | ···················· | ···················· | ···················· | ···················· | ···················· | ···················· |

| | | | | | | | | |
|---|---|---|---|---|---|---|---|---|
| A | 1675 | hCoV-19/England/20129037304/2020\|EPI_ISL_424038\|20 | ···················· | ···················· | ···················· | ···················· | ···················· | ···················· |
| A | 1677 | hCoV-19/England/20129059204/2020\|EPI_ISL_424054\|20 | ···················· | ···················· | ···················· | ···················· | ···················· | ···················· |
| A | 1679 | hCoV-19/England/20129085704/2020\|EPI_ISL_424076\|20 | ···················· | ···················· | ···················· | ···················· | ···················· | ···················· |
| A | 1680 | hCoV-19/England/20129089504/2020\|EPI_ISL_424078\|20 | ···················· | ···················· | ···················· | ···················· | ···················· | ···················· |
| A | 1681 | hCoV-19/England/20129096604/2020\|EPI_ISL_424082\|20 | ···················· | ···················· | ···················· | ···················· | ···················· | ···················· |
| A | 1682 | hCoV-19/England/20129096804/2020\|EPI_ISL_424084\|20 | ···················· | ···················· | ···················· | ···················· | ···················· | ···················· |
| A | 1684 | hCoV-19/England/CAMB-74571/2020\|EPI_ISL_425307\|202 | ···················· | ···················· | ···················· | ···················· | ···················· | ···················· |
| A | 1685 | hCoV-19/England/CAMB-74580/2020\|EPI_ISL_425308\|202 | ···················· | ···················· | ···················· | ···················· | ···················· | ···················· |
| A | 1686 | hCoV-19/England/CAMB-7459F/2020\|EPI_ISL_425309\|202 | ···················· | ···················· | ···················· | ···················· | ···················· | ···················· |
| A | 1687 | hCoV-19/England/CAMB-745AE/2020\|EPI_ISL_425310\|202 | ···················· | ···················· | ···················· | ···················· | ···················· | ···················· |
| A | 1688 | hCoV-19/England/CAMB-745BD/2020\|EPI_ISL_425311\|202 | ···················· | ···················· | ···················· | ···················· | ···················· | ···················· |
| A | 1689 | hCoV-19/England/CAMB-745CC/2020\|EPI_ISL_425312\|202 | ···················· | ···················· | ···················· | ···················· | ···················· | ···················· |
| A | 1690 | hCoV-19/England/CAMB-745DB/2020\|EPI_ISL_425313\|202 | ···················· | ···················· | ···················· | ···················· | ···················· | ···················· |
| A | 1691 | hCoV-19/England/CAMB-745EA/2020\|EPI_ISL_425314\|202 | ···················· | ···················· | ···················· | ···················· | ···················· | ···················· |
| A | 1692 | hCoV-19/England/CAMB-74605/2020\|EPI_ISL_425315\|202 | ···················· | ···················· | ···················· | ···················· | ···················· | ···················· |
| A | 1693 | hCoV-19/England/CAMB-74614/2020\|EPI_ISL_425316\|202 | ···················· | ···················· | ···················· | ···················· | ···················· | ···················· |
| A | 1694 | hCoV-19/England/CAMB-74623/2020\|EPI_ISL_425317\|202 | ···················· | ···················· | ···················· | ···················· | ···················· | ···················· |
| A | 1695 | hCoV-19/England/CAMB-74641/2020\|EPI_ISL_425318\|202 | ···················· | ···················· | ···················· | ···················· | ···················· | ···················· |
| A | 1696 | hCoV-19/England/CAMB-74650/2020\|EPI_ISL_425319\|202 | ···················· | ···················· | ···················· | ···················· | ···················· | ···················· |
| A | 1700 | hCoV-19/England/CAMB-746BA/2020\|EPI_ISL_425324\|202 | ···················· | ···················· | ···················· | ···················· | ···················· | ···················· |
| A | 1701 | hCoV-19/England/CAMB-746D8/2020\|EPI_ISL_425325\|202 | ···················· | ···················· | ···················· | ···················· | ···················· | ···················· |
| A | 1702 | hCoV-19/England/CAMB-74702/2020\|EPI_ISL_425326\|202 | ···················· | ···················· | ···················· | ···················· | ···················· | ···················· |
| A | 1703 | hCoV-19/England/CAMB-74711/2020\|EPI_ISL_425327\|202 | ···················· | ···················· | ···················· | ···················· | ···················· | ···················· |
| A | 1704 | hCoV-19/England/CAMB-7475D/2020\|EPI_ISL_425331\|202 | ···················· | ···················· | ···················· | ···················· | ···················· | ···················· |
| A | 1705 | hCoV-19/England/CAMB-7476C/2020\|EPI_ISL_425332\|202 | ···················· | ···················· | ···················· | ···················· | ···················· | ···················· |
| A | 1706 | hCoV-19/England/CAMB-7477B/2020\|EPI_ISL_425333\|202 | ···T················ | ···················· | ···················· | ···················· | ···················· | ···················· |
| A | 1707 | hCoV-19/England/CAMB-748E1/2020\|EPI_ISL_425335\|202 | ···················· | ···················· | ···················· | ···················· | ···················· | ···················· |
| A | 1708 | hCoV-19/England/CAMB-7491B/2020\|EPI_ISL_425338\|202 | ···················· | ···················· | ···················· | ···················· | ···················· | ···················· |
| A | 1709 | hCoV-19/England/CAMB-7492A/2020\|EPI_ISL_425339\|202 | ···················· | ···················· | ···················· | ···················· | ···················· | ···················· |
| A | 1711 | hCoV-19/England/CAMB-74B42/2020\|EPI_ISL_425348\|202 | ···················· | ···················· | ···················· | ···················· | ···················· | NN·················· |
| A | 1715 | hCoV-19/England/CAMB-74C30/2020\|EPI_ISL_425359\|202 | ···················· | ···················· | ···················· | ···················· | ···················· | ···················· |
| A | 1716 | hCoV-19/England/CAMB-74C5E/2020\|EPI_ISL_425361\|202 | ···················· | ···················· | ···················· | ···················· | ···················· | ···················· |
| A | 1717 | hCoV-19/England/CAMB-74C8B/2020\|EPI_ISL_425362\|202 | ···················· | ···················· | ···················· | ···················· | ···················· | ···················· |
| A | 1718 | hCoV-19/England/CAMB-74D5B/2020\|EPI_ISL_425373\|202 | ···················· | ···················· | ···················· | ···················· | ···················· | ···················· |
| A | 1719 | hCoV-19/England/CAMB-74D6A/2020\|EPI_ISL_425374\|202 | ···················· | ···················· | ···················· | ···················· | ···················· | ···················· |
| A | 1720 | hCoV-19/England/CAMB-74E49/2020\|EPI_ISL_425378\|202 | ···················· | ···················· | ···················· | ···················· | ···················· | ···················· |
| A | 1721 | hCoV-19/England/CAMB-74E67/2020\|EPI_ISL_425380\|202 | ···················· | ···················· | ···················· | ···················· | ···················· | ···················· |
| A | 1722 | hCoV-19/England/CAMB-74E85/2020\|EPI_ISL_425382\|202 | ···················· | ···················· | ···················· | ···················· | ···················· | ···················· |
| A | 1728 | hCoV-19/Australia/VIC327/2020\|EPI_ISL_426637\|2020- | ···················· | ···················· | ···················· | ···················· | ···················· | ···················· |
| A | 1729 | hCoV-19/Australia/VIC328/2020\|EPI_ISL_426638\|2020- | ···················· | ···················· | ···················· | ···················· | ···················· | ···················· |
| A | 1730 | hCoV-19/Australia/VIC329/2020\|EPI_ISL_426639\|2020- | ···················· | ···················· | ···················· | ···················· | ···················· | ···················· |
| A | 1731 | hCoV-19/Australia/VIC330/2020\|EPI_ISL_426640\|2020- | ···················· | ···················· | ···················· | ···················· | ···················· | ···················· |
| A | 1732 | hCoV-19/Australia/VIC332/2020\|EPI_ISL_426641\|2020- | ···················· | ···················· | ···················· | ···················· | ···················· | ···················· |
| A | 1733 | hCoV-19/Australia/VIC333/2020\|EPI_ISL_426642\|2020- | ···················· | ···················· | ···················· | ···················· | ···················· | ···················· |
| A | 1734 | hCoV-19/Australia/VIC334/2020\|EPI_ISL_426643\|2020- | ···················· | ···················· | ···················· | ···················· | ···················· | ···················· |
| A | 1735 | hCoV-19/Australia/VIC335/2020\|EPI_ISL_426644\|2020- | ···················· | ···················· | ···················· | ···················· | ···················· | ···················· |
| A | 1736 | hCoV-19/Australia/VIC336/2020\|EPI_ISL_426645\|2020- | ···················· | ···················· | ···················· | ···················· | ···················· | ···················· |
| A | 1737 | hCoV-19/Australia/VIC337/2020\|EPI_ISL_426646\|2020- | ···················· | ···················· | ···················· | ···················· | ···················· | ···················· |
| A | 1738 | hCoV-19/Australia/VIC338/2020\|EPI_ISL_426647\|2020- | ···················· | ···················· | ···················· | ···················· | ···················· | ···················· |
| A | 1739 | hCoV-19/Australia/VIC339/2020\|EPI_ISL_426648\|2020- | ···················· | ···················· | ···················· | ···················· | ···················· | ···················· |
| A | 1740 | hCoV-19/Australia/VIC340/2020\|EPI_ISL_426649\|2020- | ···················· | ···················· | ···················· | ···················· | ···················· | ···················· |
| A | 1741 | hCoV-19/Australia/VIC341/2020\|EPI_ISL_426650\|2020- | ···················· | ·A·················· | ···················· | ···················· | ···················· | ···················· |
| A | 1742 | hCoV-19/Australia/VIC342/2020\|EPI_ISL_426651\|2020- | ···················· | ···················· | ···················· | ···················· | ···················· | ···················· |
| A | 1743 | hCoV-19/Australia/VIC343/2020\|EPI_ISL_426652\|2020- | ···················· | ···················· | ···················· | ···················· | ···················· | ···················· |
| A | 1744 | hCoV-19/Australia/VIC344/2020\|EPI_ISL_426653\|2020- | ···················· | ···················· | ···················· | ···················· | ···················· | ···················· |
| A | 1745 | hCoV-19/Australia/VIC345/2020\|EPI_ISL_426654\|2020- | ···················· | ···················· | ···················· | ···················· | ···················· | ···················· |
| A | 1746 | hCoV-19/Australia/VIC346/2020\|EPI_ISL_426655\|2020- | ···················· | ···················· | ···················· | ···················· | ···················· | ···················· |
| A | 1747 | hCoV-19/Australia/VIC347/2020\|EPI_ISL_426656\|2020- | ···················· | ···················· | ···················· | ···················· | ···················· | ···················· |
| A | 1748 | hCoV-19/Australia/VIC348/2020\|EPI_ISL_426657\|2020- | ···················· | ···················· | ···················· | ···················· | ···················· | ···················· |
| A | 1749 | hCoV-19/Australia/VIC350/2020\|EPI_ISL_426658\|2020- | ···················· | ···················· | ···················· | ···················· | ···················· | ···················· |
| A | 1750 | hCoV-19/Australia/VIC351/2020\|EPI_ISL_426659\|2020- | ···················· | ···················· | ···················· | ···················· | ···················· | ···················· |
| A | 1751 | hCoV-19/Australia/VIC353/2020\|EPI_ISL_426661\|2020- | ···················· | ···················· | ···················· | ···················· | ···················· | ···················· |
| A | 1752 | hCoV-19/Australia/VIC354/2020\|EPI_ISL_426662\|2020- | ···················· | ···················· | ···················· | ···················· | ···················· | ···················· |
| A | 1753 | hCoV-19/Australia/VIC355/2020\|EPI_ISL_426663\|2020- | ···················· | ···················· | ···················· | ···················· | ···················· | ···················· |
| A | 1754 | hCoV-19/Australia/VIC356/2020\|EPI_ISL_426664\|2020- | ···················· | ···················· | ···················· | ···················· | ···················· | ···················· |
| A | 1755 | hCoV-19/Australia/VIC357/2020\|EPI_ISL_426665\|2020- | ···················· | ···················· | ···················· | ···················· | ···················· | ···················· |
| A | 1756 | hCoV-19/Australia/VIC358/2020\|EPI_ISL_426666\|2020- | ···················· | ···················· | ···················· | ···················· | ···················· | ···················· |
| A | 1757 | hCoV-19/Australia/VIC359/2020\|EPI_ISL_426667\|2020- | ···················· | ···················· | ···················· | ···················· | ···················· | ···················· |
| A | 1758 | hCoV-19/Australia/VIC361/2020\|EPI_ISL_426669\|2020- | ···················· | ···················· | ···················· | ···················· | ···················· | ···················· |
| A | 1759 | hCoV-19/Australia/VIC362/2020\|EPI_ISL_426670\|2020- | ···················· | ···················· | ···················· | ···················· | ···················· | ···················· |
| A | 1760 | hCoV-19/Australia/VIC363/2020\|EPI_ISL_426671\|2020- | ···················· | ···················· | ···················· | ···················· | ···················· | ···················· |
| A | 1761 | hCoV-19/Australia/VIC364/2020\|EPI_ISL_426672\|2020- | ···················· | ···················· | ···················· | ···················· | ···················· | ···················· |
| A | 1762 | hCoV-19/Australia/VIC365/2020\|EPI_ISL_426673\|2020- | ···················· | ···················· | ···················· | ···················· | ···················· | ···················· |
| A | 1763 | hCoV-19/Australia/VIC366/2020\|EPI_ISL_426674\|2020- | ···················· | ···················· | ···················· | ···················· | ···················· | ···················· |
| A | 1764 | hCoV-19/Australia/VIC367/2020\|EPI_ISL_426675\|2020- | ···················· | ···················· | ···················· | ···················· | ···················· | ···················· |
| A | 1765 | hCoV-19/Australia/VIC368/2020\|EPI_ISL_426676\|2020- | ···················· | ···················· | ···················· | ···················· | ···················· | ···················· |
| A | 1766 | hCoV-19/Australia/VIC369/2020\|EPI_ISL_426677\|2020- | ···················· | ···················· | ···················· | ···················· | ···················· | ···················· |
| A | 1767 | hCoV-19/Australia/VIC372/2020\|EPI_ISL_426679\|2020- | ···················· | ···················· | ···················· | ···················· | ···················· | ···················· |

```
A    1768 hCoV-19/Australia/VIC373/2020|EPI_ISL_426680|2020-   ....................  ....................  ....................  ....................  ....................  ....................
A    1769 hCoV-19/Australia/VIC375/2020|EPI_ISL_426682|2020-   ....................  ....................  ....................  ....................  ....................  ....................
A    1770 hCoV-19/Australia/VIC376/2020|EPI_ISL_426683|2020-   ....................  ....................  ....................  ....................  ....................  ....................
A    1771 hCoV-19/Australia/VIC377/2020|EPI_ISL_426684|2020-   ....................  ....................  ....................  ....................  ....................  ....................
A    1772 hCoV-19/Australia/VIC378/2020|EPI_ISL_426685|2020-   ....................  ....................  ....................  ....................  ....................  ....................
A    1773 hCoV-19/Australia/VIC379/2020|EPI_ISL_426686|2020-   ....................  ....................  ....................  ....................  ....................  ....................
A    1774 hCoV-19/Australia/VIC380/2020|EPI_ISL_426687|2020-   ....................  ....................  ....................  ....................  ....................  ....................
A    1775 hCoV-19/Australia/VIC381/2020|EPI_ISL_426688|2020-   ....................  ....................  ....................  ....................  ....................  ....................
A    1776 hCoV-19/Australia/VIC382/2020|EPI_ISL_426689|2020-   ....................  ....................  ....................  ....................  ....................  ....................
A    1777 hCoV-19/Australia/VIC383/2020|EPI_ISL_426690|2020-   ....................  ....................  ....................  ....................  ....................  ....................
A    1778 hCoV-19/Australia/VIC384/2020|EPI_ISL_426691|2020-   ....................  ....................  ....................  ....................  ....................  ....................
A    1779 hCoV-19/Australia/VIC385/2020|EPI_ISL_426692|2020-   ....................  ....................  ....................  ....................  ....................  ....................
A    1780 hCoV-19/Australia/VIC386/2020|EPI_ISL_426693|2020-   ....................  ....................  ....................  ....................  ....................  ....................
A    1781 hCoV-19/Australia/VIC387/2020|EPI_ISL_426694|2020-   ....................  ....................  ....................  ....................  ....................  ....................
A    1782 hCoV-19/Australia/VIC389/2020|EPI_ISL_426696|2020-   ....................  ....................  ....................  ....................  ....................  ....................
A    1783 hCoV-19/Australia/VIC394/2020|EPI_ISL_426697|2020-   ....................  ....................  ....................  ....................  ....................  ....................
A    1784 hCoV-19/Australia/VIC395/2020|EPI_ISL_426698|2020-   ....................  ....................  ....................  ....................  ....................  ....................
A    1785 hCoV-19/Australia/VIC396/2020|EPI_ISL_426699|2020-   ....................  ....................  ....................  ....................  ....................  ....................
A    1786 hCoV-19/India/DL-NIV-763/2020|EPI_ISL_547870|2020    ....................  ....................  ....................  ....................  ....................  ....................
A    1787 hCoV-19/India/DL-NIV-770/2020|EPI_ISL_547871|2020    ....................  ....................  ....................  ....................  ....................  ....................
A    1788 hCoV-19/India/DL-NIV-772/2020|EPI_ISL_547872|2020    ....................  ....................  ....................  ....................  ....................  ....................
A    1789 hCoV-19/India/DL-NIV-773/2020|EPI_ISL_547873|2020    ....................  ....................  ....................  ....................  ....................  ....................
A    1790 hCoV-19/India/DL-NIV-777/2020|EPI_ISL_547874|2020    ....................  ....................  ....................  ....................  ....................  ....................
A    1791 hCoV-19/India/DL-NIV-781/2020|EPI_ISL_547875|2020    ....................  ....................  ....................  ....................  ....................  ....................
A    1792 hCoV-19/India/UP-NIV-32/2020|EPI_ISL_547876|2020     ....................  ....................  ....................  ....................  ....................  ....................
A    1793 hCoV-19/Wuhan/IPBCAMS-WH-03/2019|EPI_ISL_403930|20   ....................  ....................  ....................  ....................  ....................  ....................
A    1794 hCoV-19/Wuhan/IPBCAMS-WH-02/2019|EPI_ISL_403931|20   ....................  ....................  ....................  ....................  ....................  ....................
A    1795 hCoV-19/Guangdong/20SF012/2020|EPI_ISL_403932|2020   ....................  ....................  ....................  ....................  ....................  ....................
A    1796 hCoV-19/Thailand/61/2020|EPI_ISL_403962|2020-01-08   ....................  ....................  ....................  ....................  ....................  ....................
A    1797 hCoV-19/Thailand/74/2020|EPI_ISL_403963|2020-01-13   ....................  ....................  ....................  ....................  ....................  ....................
A    1798 hCoV-19/France/GES-1583/2020|EPI_ISL_414600|2020-0   ....................  ....................  ....................  ....................  ....................  ....................
A    1799 hCoV-19/USA/WA-UW29/2020|EPI_ISL_414616|2020-03-08   ....................  ....................  ....................  ....................  ....................  ....................
A    1800 hCoV-19/USA/WA-UW30/2020|EPI_ISL_414617|2020-03-08   ....................  ....................  ....................  ....................  ....................  ....................
A    1801 hCoV-19/USA/WA-UW31/2020|EPI_ISL_414618|2020-03-08   ....................  ....................  ....................  ....................  ....................  ....................
A    1802 hCoV-19/USA/WA-UW32/2020|EPI_ISL_414619|2020-03-07   ....................  ....................  ....................  ....................  ....................  ....................
A    1803 hCoV-19/USA/WA-UW33/2020|EPI_ISL_414620|2020-03-08   ....................  ....................  ....................  ....................  ....................  ....................
A    1804 hCoV-19/USA/WA-UW34/2020|EPI_ISL_414621|2020-03-07   ....................  ....................  ....................  ....................  ....................  ....................
A    1805 hCoV-19/USA/WA-UW35/2020|EPI_ISL_414622|2020-03-08   ....................  ....................  ....................  ....................  ....................  ....................
A    1806 hCoV-19/France/GES-1583-2/2020|EPI_ISL_414623|2020   ....................  ....................  ....................  ....................  ....................  ....................
A    1807 hCoV-19/France/NOR-N1620/2020|EPI_ISL_414624|2020-   ....................  ....................  ....................  ....................  ....................  ....................
A    1808 hCoV-19/France/PDL-1643/2020|EPI_ISL_414625|2020-0   ....................  ....................  ....................  ....................  ....................  ....................
A    1809 hCoV-19/France/HDF-1684/2020|EPI_ISL_414626|2020-0   ....................  ....................  ....................  ....................  ....................  ....................
A    1810 hCoV-19/France/HDF-1795/2020|EPI_ISL_414627|2020-0   ....................  ....................  ....................  ....................  ....................  ....................
A    1811 hCoV-19/France/HDF-1805/2020|EPI_ISL_414628|2020-0   ....................  ....................  ....................  ....................  ....................  ....................
A    1812 hCoV-19/France/HDF-1870/2020|EPI_ISL_414629|2020-0   ....................  ....................  ....................  ....................  ....................  ....................
A    1813 hCoV-19/France/HDF-1871/2020|EPI_ISL_414630|2020-0   ....................  ....................  ....................  ....................  ....................  ....................
A    1814 hCoV-19/France/GES-1973/2020|EPI_ISL_414631|2020-0   ....................  ....................  ....................  ....................  ....................  ....................
A    1815 hCoV-19/France/GES-1977/2020|EPI_ISL_414632|2020-0   ....................  ....................  ....................  ....................  ....................  ....................
A    1816 hCoV-19/France/IDF-1980/2020|EPI_ISL_414633|2020-0   ....................  ....................  ....................  ....................  ....................  ....................
A    1817 hCoV-19/France/HDF-1986/2020|EPI_ISL_414634|2020-0   ....................  ....................  ....................  ....................  ....................  ....................
A    1818 hCoV-19/France/HDF-1988/2020|EPI_ISL_414635|2020-0   ....................  ....................  ....................  ....................  ....................  ....................
A    1819 hCoV-19/France/HDF-1989/2020|EPI_ISL_414636|2020-0   ....................  ....................  ....................  ....................  ....................  ....................
A    1820 hCoV-19/France/HDF-1993/2020|EPI_ISL_414637|2020-0   ....................  ....................  ....................  ....................  ....................  ....................
A    1821 hCoV-19/France/HDF-1995/2020|EPI_ISL_414638|2020-0   ....................  ....................  ....................  ....................  ....................  ....................
A    1822 hCoV-19/Finland/FIN-313/2020|EPI_ISL_414641|2020-0   ....................  ....................  ....................  ....................  ....................  ....................
A    1823 hCoV-19/Finland/FIN-455/2020|EPI_ISL_414642|2020-0   ....................  ....................  ....................  ....................  ....................  ....................
A    1824 hCoV-19/Finland/FIN-508/2020|EPI_ISL_414643|2020-0   ....................  ....................  ....................  ....................  ....................  ....................
A    1825 hCoV-19/Finland/FIN-266/2020|EPI_ISL_414646|2020-0   ....................  ....................  ....................  ....................  ....................  ....................
A    1826 hCoV-19/USA/CA-PC101P/2020|EPI_ISL_414648|2020-03-   ....................  ....................  ....................  ....................  ....................  ....................
A    1827 hCoV-19/Spain/MD-ISCIII-201442/2020|EPI_ISL_417010   ....................  ....................  ....................  ....................  ....................  ....................
A    1830 hCoV-19/Belgium/ULG-6216/2020|EPI_ISL_417014|2020-   ....................  ....................  ....................  ....................  ....................  ....................
A    1831 hCoV-19/Belgium/ULG-6457/2020|EPI_ISL_417015|2020-   ....................  ....................  ....................  ....................  ....................  ....................
A    1832 hCoV-19/Belgium/ULG-6503/2020|EPI_ISL_417016|2020-   ....................  ....................  ....................  ....................  ....................  ....................
A    1833 hCoV-19/Belgium/ULG-6638/2020|EPI_ISL_417017|2020-   ....................  ....................  ....................  ....................  ....................  ....................
A    1834 hCoV-19/Belgium/ULG-6670/2020|EPI_ISL_417018|2020-   ....................  ....................  ....................  ....................  ....................  ....................
A    1835 hCoV-19/Belgium/ULG-6754/2020|EPI_ISL_417019|2020-   ....................  ....................  ....................  ....................  ....................  ....................
A    1836 hCoV-19/Belgium/ULG-6939/2020|EPI_ISL_417020|2020-   ....................  ....................  ....................  ....................  ....................  ....................
A    1837 hCoV-19/Belgium/ULG-6942/2020|EPI_ISL_417021|2020-   ....................  ..........A.........  ....................  ....................  ....................  ....................
A    1838 hCoV-19/Belgium/ULG-6948/2020|EPI_ISL_417022|2020-   ....................  ..........A.........  ....................  ....................  ....................  ....................
A    1839 hCoV-19/Belgium/ULG-6950/2020|EPI_ISL_417023|2020-   ....................  ..........A.........  ....................  ....................  ....................  ....................
A    1840 hCoV-19/Belgium/ULG-7019/2020|EPI_ISL_417025|2020-   ....................  ....................  ....................  ....................  ....................  ....................
A    1841 hCoV-19/USA/UT-08/2020|EPI_ISL_417026|2020-03-20     ....................  ....................  ....................  ....................  ....................  ....................
A    1842 hCoV-19/USA/UT-09/2020|EPI_ISL_417027|2020-03-20     ....................  ....................  ....................  ....................  ....................  ....................
A    1843 hCoV-19/USA/UT-020/2020|EPI_ISL_417028|2020-03-20    ....................  ....................  ....................  ....................  ....................  ....................
A    1844 hCoV-19/Australia/NSW04/2020|EPI_ISL_417030|2020-0   ....................  ....................  ....................  ....................  ....................  ....................
A    1845 hCoV-19/Australia/QLDID919/2020|EPI_ISL_417031|202   ....................  ....................  ....................  ....................  ....................  ....................
A    1846 hCoV-19/Australia/QLDID920/2020|EPI_ISL_417032|202   ....................  ....................  ....................  ....................  ....................  ....................
A    1847 hCoV-19/Australia/QLDID921/2020|EPI_ISL_417033|202   ....................  ....................  ....................  ....................  ....................  ....................
```

| | | | | | | | | |
|---|---|---|---|---|---|---|---|---|
| A | 1848 | hCoV-19/Brazil/AM-02/2020\|EPI_ISL_417034\|2020-03-1 | ···················· | ···················· | ····················· | ···················· | ···················· | ···················· |
| A | 1849 | hCoV-19/Hong Kong/VB20017970-2/2020\|EPI_ISL_417064 | ···················· | ···················· | ····················· | ···················· | ···················· | ···················· |
| A | 1850 | hCoV-19/USA/WA-S12/2020\|EPI_ISL_417065\|2020-03-03 | ···················· | ···················· | ····················· | ···················· | ···················· | ···················· |
| A | 1851 | hCoV-19/USA/WA-S13/2020\|EPI_ISL_417066\|2020-03-03 | ···················· | ···················· | ····················· | ···················· | ···················· | ···················· |
| A | 1852 | hCoV-19/USA/WA-S15/2020\|EPI_ISL_417068\|2020-03-02 | ···················· | ···················· | ····················· | ···················· | ···················· | ···················· |
| A | 1853 | hCoV-19/USA/WA-S16/2020\|EPI_ISL_417069\|2020-03-03 | ···················· | ···················· | ····················· | ···················· | ···················· | ···················· |
| A | 1854 | hCoV-19/USA/WA-S32/2020\|EPI_ISL_417085\|2020-03-02 | ···················· | ···················· | ····················· | ···················· | ···················· | ···················· |
| A | 1855 | hCoV-19/USA/WA-S33/2020\|EPI_ISL_417086\|2020-03-01 | ···················· | ···················· | ····················· | ···················· | ···················· | ···················· |
| A | 1856 | hCoV-19/USA/WA-S34/2020\|EPI_ISL_417087\|2020-03-02 | ···················· | ···················· | ····················· | ···················· | ···················· | ···················· |
| A | 1857 | hCoV-19/USA/WA-S35/2020\|EPI_ISL_417088\|2020-03-02 | ···················· | ···················· | ····················· | ···················· | ···················· | ···················· |
| A | 1858 | hCoV-19/USA/WA-S36/2020\|EPI_ISL_417089\|2020-03-02 | ···················· | ···················· | ····················· | ···················· | ···················· | ···················· |
| A | 1859 | hCoV-19/Hungary/mbl2/2020\|EPI_ISL_418183\|2020-03-1 | ···················· | ···················· | ····················· | ···················· | ···················· | ···················· |
| A | 1860 | hCoV-19/Canada/ON-PHL-1083/2020\|EPI_ISL_418340\|202 | ···················· | ···················· | ····················· | ···················· | ···················· | ···················· |
| A | 1861 | hCoV-19/Canada/ON-PHL-0142/2020\|EPI_ISL_418341\|202 | ···················· | ···················· | ····················· | ···················· | ···················· | ···················· |
| A | 1862 | hCoV-19/Canada/ON-PHL-0178/2020\|EPI_ISL_418342\|202 | ···················· | ···················· | ····················· | ···················· | ···················· | ···················· |
| A | 1863 | hCoV-19/Canada/ON-PHL-6884/2020\|EPI_ISL_418343\|202 | ···················· | ···················· | ····················· | ···················· | ···················· | ···················· |
| A | 1864 | hCoV-19/Canada/ON-PHL-2259/2020\|EPI_ISL_418344\|202 | ···················· | ···················· | ····················· | ···················· | ···················· | ···················· |
| A | 1865 | hCoV-19/Canada/ON-PHL-8751/2020\|EPI_ISL_418345\|202 | ···················· | ···················· | ····················· | ···················· | ···················· | ···················· |
| A | 1866 | hCoV-19/Canada/ON-PHL-0743/2020\|EPI_ISL_418346\|202 | ···················· | ···················· | ····················· | ···················· | ···················· | ···················· |
| A | 1867 | hCoV-19/Canada/ON-PHL-3741/2020\|EPI_ISL_418347\|202 | ···················· | ···················· | ····················· | ···················· | ···················· | ···················· |
| A | 1868 | hCoV-19/Canada/ON-PHL-3680/2020\|EPI_ISL_418348\|202 | ···················· | ···················· | ····················· | ···················· | ···················· | ···················· |
| A | 1869 | hCoV-19/Canada/ON-PHL-3650/2020\|EPI_ISL_418349\|202 | ···················· | ···················· | ····················· | ···················· | ···················· | ···················· |
| A | 1870 | hCoV-19/Canada/ON-PHL-3380/2020\|EPI_ISL_418350\|202 | ···················· | ···················· | ····················· | ···················· | ···················· | ···················· |
| A | 1871 | hCoV-19/Canada/ON-PHL-3575/2020\|EPI_ISL_418351\|202 | ···················· | ···················· | ····················· | ···················· | ···················· | ···················· |
| A | 1872 | hCoV-19/Canada/ON-PHL-5672/2020\|EPI_ISL_418352\|202 | ···················· | ···················· | ····················· | ···················· | ···················· | ···················· |
| A | 1873 | hCoV-19/Canada/ON-PHL-0539/2020\|EPI_ISL_418353\|202 | ···················· | ···················· | ····················· | ···················· | ···················· | ···················· |
| A | 1874 | hCoV-19/Canada/ON-PHL-4088/2020\|EPI_ISL_418354\|202 | ···················· | ···················· | ····················· | ···················· | ···················· | ···················· |
| A | 1875 | hCoV-19/Canada/ON-PHL-U8150/2020\|EPI_ISL_418355\|20 | ···················· | ···················· | ····················· | ···················· | ···················· | ···················· |
| A | 1876 | hCoV-19/Canada/ON-PHL-6980/2020\|EPI_ISL_418356\|202 | ···················· | ···················· | ····················· | ···················· | ···················· | ···················· |
| A | 1877 | hCoV-19/Canada/ON-PHL-H6415/2020\|EPI_ISL_418357\|20 | ···················· | ···················· | ····················· | ···················· | ···················· | ···················· |
| A | 1878 | hCoV-19/Canada/ON-PHL-5756/2020\|EPI_ISL_418358\|202 | ···················· | ···················· | ····················· | ···················· | ···················· | ··A················· |
| A | 1879 | hCoV-19/Canada/ON-PHL-5757/2020\|EPI_ISL_418359\|202 | ···················· | ···················· | ····················· | ···················· | ···················· | ··A················· |
| A | 1880 | hCoV-19/Canada/ON-PHL-3692/2020\|EPI_ISL_418360\|202 | ···················· | ···················· | ····················· | ···················· | ···················· | ···················· |
| A | 1881 | hCoV-19/Canada/ON-PHL-1095/2020\|EPI_ISL_418361\|202 | ···················· | ···················· | ····················· | ···················· | ···················· | ···················· |
| A | 1882 | hCoV-19/Canada/ON-PHL-3536/2020\|EPI_ISL_418362\|202 | ···················· | ···················· | ····················· | ···················· | ···················· | ···················· |
| A | 1883 | hCoV-19/Canada/ON-PHL-3695/2020\|EPI_ISL_418363\|202 | ···················· | ···················· | ····················· | ···················· | ···················· | ···················· |
| A | 1884 | hCoV-19/Canada/ON-PHL-6922/2020\|EPI_ISL_418364\|202 | ···················· | ···················· | ····················· | ···················· | ···················· | ···················· |
| A | 1885 | hCoV-19/Canada/ON-PHL-0977/2020\|EPI_ISL_418365\|202 | ···················· | ···················· | ····················· | ···················· | ···················· | ···················· |
| A | 1886 | hCoV-19/Canada/ON-PHL-3350/2020\|EPI_ISL_418366\|202 | ···················· | ···················· | ····················· | ···················· | ···················· | ···················· |
| A | 1887 | hCoV-19/Canada/ON-PHL-7513/2020\|EPI_ISL_418367\|202 | ···················· | ···················· | ····················· | ···················· | ···················· | ···················· |
| A | 1888 | hCoV-19/Canada/ON-PHL-3458/2020\|EPI_ISL_418368\|202 | ···················· | ···················· | ····················· | ···················· | ···················· | ···················· |
| A | 1889 | hCoV-19/Canada/ON-PHL-8539/2020\|EPI_ISL_418369\|202 | ···················· | ···················· | ····················· | ···················· | ···················· | ···················· |
| A | 1890 | hCoV-19/Canada/ON-PHL-0654/2020\|EPI_ISL_418370\|202 | ···················· | ···················· | ····················· | ···················· | ···················· | ···················· |
| A | 1891 | hCoV-19/Canada/ON-PHL-2653/2020\|EPI_ISL_418371\|202 | ···················· | ···················· | ····················· | ···················· | ···················· | ···················· |
| A | 1892 | hCoV-19/Canada/ON-PHL-8458/2020\|EPI_ISL_418372\|202 | ···················· | ···················· | ····················· | ···················· | ···················· | ···················· |
| A | 1893 | hCoV-19/Canada/ON-PHL-7590/2020\|EPI_ISL_418373\|202 | ···················· | ···················· | ····················· | ···················· | ···················· | ···················· |
| A | 1894 | hCoV-19/Canada/ON-PHL-4232/2020\|EPI_ISL_418374\|202 | ···················· | ···················· | ····················· | ···················· | ···················· | ···················· |
| A | 1895 | hCoV-19/Canada/ON-PHL-0141/2020\|EPI_ISL_418375\|202 | ···················· | ···················· | ····················· | ···················· | ···················· | ···················· |
| A | 1896 | hCoV-19/Canada/ON-PHL-3459/2020\|EPI_ISL_418376\|202 | ···················· | ···················· | ····················· | ···················· | ···················· | ···················· |
| A | 1897 | hCoV-19/Canada/ON-PHL-1898/2020\|EPI_ISL_418377\|202 | ···················· | ···················· | ····················· | ···················· | ···················· | ···················· |
| A | 1898 | hCoV-19/Canada/ON-PHL-0976/2020\|EPI_ISL_418378\|202 | ···················· | ···················· | ····················· | ···················· | ···················· | ···················· |
| A | 1899 | hCoV-19/Canada/ON-PHL-7512/2020\|EPI_ISL_418379\|202 | ···················· | ···················· | ····················· | ···················· | ···················· | ···················· |
| A | 1900 | hCoV-19/Canada/ON-PHL-3476/2020\|EPI_ISL_418380\|202 | ···················· | ···················· | ····················· | ···················· | ···················· | ···················· |
| A | 1901 | hCoV-19/Canada/ON-PHL-2223/2020\|EPI_ISL_418381\|202 | ···················· | ···················· | ····················· | ···················· | ···················· | ···················· |
| A | 1902 | hCoV-19/Canada/ON-PHL-5930/2020\|EPI_ISL_418382\|202 | ···················· | ···················· | ····················· | ···················· | ···················· | ···················· |
| A | 1903 | hCoV-19/Canada/ON-PHL-2273/2020\|EPI_ISL_418383\|202 | ···················· | ···················· | ····················· | ···················· | ···················· | ···················· |
| A | 1904 | hCoV-19/Finland/13M19/2020\|EPI_ISL_418385\|2020-03- | ···················· | ···················· | ····················· | ···················· | ···················· | ···················· |
| A | 1905 | hCoV-19/Finland/13M26/2020\|EPI_ISL_418386\|2020-03- | ···················· | ···················· | ····················· | ···················· | ···················· | ···················· |
| A | 1906 | hCoV-19/Finland/13M27/2020\|EPI_ISL_418387\|2020-03- | ···················· | ···················· | ····················· | ···················· | ···················· | ···················· |
| A | 1907 | hCoV-19/Finland/13M29/2020\|EPI_ISL_418388\|2020-03- | ···················· | ···················· | ····················· | ···················· | ···················· | ···················· |
| A | 1908 | hCoV-19/Finland/13M3/2020\|EPI_ISL_418389\|2020-03-1 | ···················· | ···················· | ····················· | ···················· | ···················· | ···················· |
| A | 1909 | hCoV-19/Finland/13M33/2020\|EPI_ISL_418390\|2020-03- | ···················· | ···················· | ····················· | ···················· | ···················· | ···················· |
| A | 1910 | hCoV-19/Finland/13M57/2020\|EPI_ISL_418391\|2020-03- | ···················· | ·A·················· | ····················· | ···················· | ···················· | ···················· |
| A | 1911 | hCoV-19/Finland/13M58/2020\|EPI_ISL_418392\|2020-03- | ···················· | ···················· | ····················· | ···················· | ···················· | ···················· |
| A | 1912 | hCoV-19/Finland/13M60/2020\|EPI_ISL_418393\|2020-03- | ···················· | ···················· | ····················· | ···················· | ···················· | ···················· |
| A | 1913 | hCoV-19/Finland/13M64/2020\|EPI_ISL_418394\|2020-03- | ···················· | ···················· | ····················· | ···················· | ···················· | ···················· |
| A | 1914 | hCoV-19/Finland/13M65/2020\|EPI_ISL_418395\|2020-03- | ···················· | ···················· | ····················· | ···················· | ···················· | ···················· |
| A | 1915 | hCoV-19/Finland/13M69/2020\|EPI_ISL_418396\|2020-03- | ···················· | ···················· | ····················· | ···················· | ···················· | ···················· |
| A | 1916 | hCoV-19/Finland/13M77/2020\|EPI_ISL_418397\|2020-03- | ···················· | ···················· | ····················· | ···················· | ···················· | ···················· |
| A | 1917 | hCoV-19/Finland/13M82/2020\|EPI_ISL_418399\|2020-03- | ···················· | ···················· | ····················· | ···················· | ···················· | ···················· |
| A | 1918 | hCoV-19/Austria/CeMM0017/2020\|EPI_ISL_419670\|2020- | ···················· | ···················· | ····················· | ···················· | ·········N·········· | ···················· |
| A | 1919 | hCoV-19/Austria/CeMM0018/2020\|EPI_ISL_419671\|2020- | ···················· | ···················· | ····················· | ···················· | ···················· | ···················· |
| A | 1920 | hCoV-19/Austria/CeMM0019/2020\|EPI_ISL_419672\|2020- | ···················· | ···················· | ····················· | ···················· | ···················· | ···················· |
| A | 1921 | hCoV-19/Austria/CeMM0020/2020\|EPI_ISL_419673\|2020- | ···················· | ···················· | ····················· | ···················· | ···················· | ···················· |
| A | 1922 | hCoV-19/Austria/CeMM0021/2020\|EPI_ISL_419674\|2020- | ···················· | ···················· | ····················· | ···················· | ···················· | ···················· |
| A | 1923 | hCoV-19/Spain/VC-FISABIO-11/2020\|EPI_ISL_419675\|20 | ···················· | ···················· | ····················· | ···················· | ···················· | ···················· |
| A | 1924 | hCoV-19/Spain/VC-FISABIO-12/2020\|EPI_ISL_419676\|20 | ···················· | ···················· | ····················· | ···················· | ···················· | ···················· |
| A | 1925 | hCoV-19/Spain/VC-FISABIO-13/2020\|EPI_ISL_419677\|20 | ···················· | ···················· | ····················· | ···················· | ···················· | ···················· |

| | | | | | | | | |
|---|---|---|---|---|---|---|---|---|
| A | 1926 | hCoV-19/Spain/VC-FISABIO-14/2020\|EPI_ISL_419678\|20 | ···················· | ···················· | ···················· | ···················· | ···················· | ···················· |
| A | 1927 | hCoV-19/Spain/VC-FISABIO-15/2020\|EPI_ISL_419679\|20 | ···················· | ···················· | ···················· | ···················· | ···················· | ···················· |
| A | 1928 | hCoV-19/Spain/VC-FISABIO-16/2020\|EPI_ISL_419680\|20 | ···················· | ···················· | ···················· | ···················· | ···················· | ···················· |
| A | 1929 | hCoV-19/Spain/VC-FISABIO-17/2020\|EPI_ISL_419681\|20 | ···················· | ···················· | ···················· | ···················· | ···················· | ···················· |
| A | 1930 | hCoV-19/Spain/VC-FISABIO-18/2020\|EPI_ISL_419682\|20 | ···················· | ···················· | ···················· | ···················· | ···················· | ···················· |
| A | 1931 | hCoV-19/Spain/VC-FISABIO-19/2020\|EPI_ISL_419683\|20 | ···················· | ···················· | ···················· | ···················· | ···················· | ···················· |
| A | 1932 | hCoV-19/Latvia/ChVir2025/2020\|EPI_ISL_419691\|2020- | ···················· | ···················· | ···················· | ···················· | ···················· | ···················· |
| A | 1933 | hCoV-19/Belarus/ChVir2072/2020\|EPI_ISL_419692\|2020 | ···················· | ······C············· | ···················· | ···················· | ···················· | ···················· |
| A | 1934 | hCoV-19/Belarus/ChVir2073/2020\|EPI_ISL_419693\|2020 | ···················· | ···················· | ···················· | ···················· | ···················· | ···················· |
| A | 1935 | hCoV-19/USA/NY-NYUMC35/2020\|EPI_ISL_419696\|2020-03 | ···················· | ···················· | ···················· | ···················· | ···················· | ···················· |
| A | 1936 | hCoV-19/USA/NY-NYUMC36/2020\|EPI_ISL_419697\|2020-03 | ···················· | ···················· | ···················· | ···················· | ···················· | ···················· |
| A | 1937 | hCoV-19/USA/NY-NYUMC37/2020\|EPI_ISL_419698\|2020-03 | ···················· | ···················· | ···················· | ···················· | ···················· | ···················· |
| A | 1938 | hCoV-19/USA/NY-NYUMC38/2020\|EPI_ISL_419699\|2020-03 | ···················· | ···················· | ···················· | ···················· | ···················· | ···················· |
| A | 1939 | hCoV-19/England/20139013406/2020\|EPI_ISL_420661\|20 | ···················· | ···················· | ···················· | ···················· | ···················· | ···················· |
| A | 1940 | hCoV-19/England/20139037504/2020\|EPI_ISL_420670\|20 | ···················· | ···················· | ···················· | ···················· | ···················· | ···················· |
| A | 1941 | hCoV-19/England/20139038104/2020\|EPI_ISL_420672\|20 | ···················· | ···················· | ···················· | ···················· | ···················· | ···················· |
| A | 1942 | hCoV-19/England/20139039504/2020\|EPI_ISL_420673\|20 | ···················· | ···················· | ···················· | ···················· | ···················· | ···················· |
| A | 1945 | hCoV-19/England/20139045904/2020\|EPI_ISL_420687\|20 | ···················· | ···················· | ···················· | ···················· | ···················· | ···················· |
| A | 1949 | hCoV-19/England/20139047104/2020\|EPI_ISL_420692\|20 | ···················· | ···················· | ···················· | ···················· | ···················· | ···················· |
| A | 1950 | hCoV-19/England/20139047404/2020\|EPI_ISL_420693\|20 | ···················· | ···················· | ···················· | ···················· | ···················· | ···················· |
| A | 1951 | hCoV-19/England/20139047504/2020\|EPI_ISL_420694\|20 | ···················· | ···················· | ···················· | ···················· | ···················· | ···················· |
| A | 1952 | hCoV-19/England/20139047604/2020\|EPI_ISL_420695\|20 | ···················· | ···················· | ···················· | ···················· | ···················· | ···················· |
| A | 1953 | hCoV-19/England/20139047804/2020\|EPI_ISL_420697\|20 | ···················· | ···················· | ···················· | ···················· | ··Y················· | ···················· |
| A | 1954 | hCoV-19/England/20139048804/2020\|EPI_ISL_420698\|20 | ···················· | ···················· | ···················· | ···················· | ··N················· | ···················· |
| A | 1955 | hCoV-19/England/20146061504/2020\|EPI_ISL_421986\|20 | ···················· | ···················· | ···················· | ···················· | ···················· | ···················· |
| A | 1956 | hCoV-19/England/CAMB-74508/2020\|EPI_ISL_425300\|202 | ···················· | ···················· | ···················· | ···················· | ···················· | ···················· |
| A | 1959 | hCoV-19/England/CAMB-74544/2020\|EPI_ISL_425304\|202 | ···················· | ···················· | ···················· | ···················· | ···················· | ···················· |
| A | 1960 | hCoV-19/England/CAMB-74553/2020\|EPI_ISL_425305\|202 | ···················· | ···················· | ···················· | ···················· | ···················· | ···················· |
| A | 1961 | hCoV-19/USA/NY-NYUMC162/2020\|EPI_ISL_426617\|2020-0 | ···················· | ···················· | ···················· | ···················· | ···················· | ···················· |
| A | 1962 | hCoV-19/USA/NY-NYUMC163/2020\|EPI_ISL_426618\|2020-0 | ···················· | ···················· | ···················· | ···················· | ···················· | ···················· |
| A | 1963 | hCoV-19/USA/NY-NYUMC164/2020\|EPI_ISL_426619\|2020-0 | ···················· | ···················· | ···················· | ···················· | ···················· | ···················· |
| A | 1964 | hCoV-19/USA/NY-NYUMC165/2020\|EPI_ISL_426620\|2020-0 | ···················· | ···················· | ···················· | ···················· | ···················· | ···················· |
| A | 1965 | hCoV-19/USA/NY-NYUMC166/2020\|EPI_ISL_426621\|2020-0 | ···················· | ···················· | ···················· | ···················· | ···················· | ···················· |
| A | 1966 | hCoV-19/USA/NY-NYUMC167/2020\|EPI_ISL_426622\|2020-0 | ···················· | ···················· | ···················· | ···················· | ···················· | ···················· |
| A | 1967 | hCoV-19/USA/NY-NYUMC168/2020\|EPI_ISL_426623\|2020-0 | ···················· | ···················· | ···················· | ···················· | ···················· | ···················· |
| A | 1968 | hCoV-19/USA/NY-NYUMC169/2020\|EPI_ISL_426624\|2020-0 | ···················· | ···················· | ···················· | ···················· | ···················· | ···················· |
| A | 1969 | hCoV-19/USA/NY-NYUMC170/2020\|EPI_ISL_426625\|2020-0 | ···················· | ···················· | ···················· | ···················· | ···················· | ···················· |
| A | 1970 | hCoV-19/USA/NY-NYUMC171/2020\|EPI_ISL_426626\|2020-0 | ···················· | ···················· | ···················· | ···················· | ···················· | ···················· |
| A | 1971 | hCoV-19/USA/LA-BIE-096/2020\|EPI_ISL_426627\|2020-04 | ···················· | ···················· | ···················· | ···················· | ···················· | ···················· |
| A | 1972 | hCoV-19/USA/LA-BIE-097/2020\|EPI_ISL_426628\|2020-04 | ···················· | ···················· | ···················· | ···················· | ···················· | ···················· |
| A | 1973 | hCoV-19/Taiwan/TSGH-01/2020\|EPI_ISL_426629\|2020-02 | ···················· | ···················· | ···················· | ···················· | ···················· | ···················· |
| A | 1974 | hCoV-19/Taiwan/TSGH-02/2020\|EPI_ISL_426630\|2020-03 | ···················· | ···················· | ···················· | ···················· | ···················· | ···················· |
| A | 1975 | hCoV-19/Taiwan/TSGH-04/2020\|EPI_ISL_426632\|2020-03 | ···················· | ···················· | ···················· | ···················· | ···················· | ···················· |
| A | 1978 | hCoV-19/Australia/NT19/2020\|EPI_ISL_426636\|2020 | ···················· | ···················· | ···················· | ···················· | ···················· | ···················· |
| A | 1979 | hCoV-19/USA/CA-CZB0103/2020\|EPI_ISL_429000\|2020-03 | ···················· | ···················· | ···················· | ···················· | ···················· | ···················· |
| A | 1980 | hCoV-19/USA/CA-CZB046/2020\|EPI_ISL_429001\|2020-03- | ···················· | ···················· | ···················· | ···················· | ···················· | ···················· |
| A | 1981 | hCoV-19/USA/CA-CZB098/2020\|EPI_ISL_429002\|2020-03- | ···················· | ···················· | ···················· | ···················· | ···················· | ···················· |
| A | 1982 | hCoV-19/USA/CA-CZB0136/2020\|EPI_ISL_429003\|2020-04 | ···················· | ···················· | ···················· | ···················· | ···················· | ···················· |
| A | 1983 | hCoV-19/USA/CA-CZB085/2020\|EPI_ISL_429004\|2020-03- | ···················· | ·A·················· | ···················· | ···················· | ···················· | ···················· |
| A | 1984 | hCoV-19/USA/CA-CZB079/2020\|EPI_ISL_429005\|2020-03- | ···················· | ···················· | ···················· | ···················· | ···················· | ···················· |
| A | 1985 | hCoV-19/USA/CA-CZB095/2020\|EPI_ISL_429006\|2020-03- | ···················· | ···················· | ···················· | ···················· | ···················· | ···················· |
| A | 1986 | hCoV-19/USA/CA-CZB0458/2020\|EPI_ISL_429007\|2020-03 | ···················· | ···················· | ···················· | ···················· | ···················· | ···················· |
| A | 1987 | hCoV-19/USA/CA-CZB091/2020\|EPI_ISL_429008\|2020-03- | ···················· | ···················· | ···················· | ···················· | ···················· | ···················· |
| A | 1988 | hCoV-19/USA/CA-CZB0122/2020\|EPI_ISL_429009\|2020-04 | ···················· | ···················· | ···················· | ···················· | ···················· | ···················· |
| A | 1989 | hCoV-19/USA/CA-CZB0108/2020\|EPI_ISL_429010\|2020-03 | ···················· | ···················· | ···················· | ···················· | ···················· | ···················· |
| A | 1990 | hCoV-19/USA/CA-CZB0147/2020\|EPI_ISL_429011\|2020-04 | ···················· | ···················· | ···················· | ···················· | ···················· | ···················· |
| A | 1991 | hCoV-19/USA/CA-CZB016/2020\|EPI_ISL_429012\|2020-03- | ···················· | ···················· | ···················· | ···················· | ···················· | ···················· |
| A | 1992 | hCoV-19/USA/CA-CZB080/2020\|EPI_ISL_429013\|2020-03- | ···················· | ···················· | ···················· | ···················· | ···················· | ···················· |
| A | 1993 | hCoV-19/USA/CA-CZB0142/2020\|EPI_ISL_429014\|2020-04 | ···················· | ···················· | ···················· | ···················· | ···················· | ···················· |
| A | 1994 | hCoV-19/USA/CA-CZB0709/2020\|EPI_ISL_429016\|2020-03 | ···················· | ····A··············· | ···················· | ···················· | ···················· | ···················· |
| A | 1995 | hCoV-19/USA/CA-CZB030/2020\|EPI_ISL_429017\|2020-03- | ···················· | ···················· | ···················· | ···················· | ···················· | ···················· |
| A | 1996 | hCoV-19/USA/CA-CZB0750/2020\|EPI_ISL_429018\|2020-04 | ···················· | ···················· | ···················· | ···················· | ···················· | ···················· |
| A | 1997 | hCoV-19/USA/CA-CZB078/2020\|EPI_ISL_429019\|2020-03- | ···················· | ···················· | ···················· | ···················· | ···················· | ···················· |
| A | 1998 | hCoV-19/USA/CA-CZB0721/2020\|EPI_ISL_429020\|2020-03 | ···················· | ···················· | ···················· | ···················· | ···················· | ···················· |
| A | 1999 | hCoV-19/USA/CA-CZB045/2020\|EPI_ISL_429021\|2020-03- | ···················· | ···················· | ···················· | ···················· | ···················· | ···················· |
| A | 2000 | hCoV-19/USA/CA-CZB0713/2020\|EPI_ISL_429022\|2020-03 | ···················· | ···················· | ···················· | ···················· | ···················· | ···················· |
| A | 2001 | hCoV-19/USA/CA-CZB0740/2020\|EPI_ISL_429023\|2020-03 | ···················· | ···················· | ···················· | ···················· | ···················· | ···················· |
| A | 2002 | hCoV-19/USA/CA-CZB0705/2020\|EPI_ISL_429024\|2020-03 | ···················· | ···················· | ···················· | ···················· | ···················· | ···················· |
| A | 2003 | hCoV-19/USA/CA-CZB0737/2020\|EPI_ISL_429025\|2020-03 | ···················· | ···················· | ···················· | ···················· | ···················· | ···················· |
| A | 2004 | hCoV-19/USA/CA-CZB0731/2020\|EPI_ISL_429026\|2020-03 | ···················· | ···················· | ···················· | ···················· | ···················· | ···················· |
| A | 2005 | hCoV-19/USA/CA-CZB0137/2020\|EPI_ISL_429027\|2020-04 | ···················· | ···················· | ···················· | ···················· | ···················· | ···················· |
| A | 2006 | hCoV-19/USA/CA-CZB0746/2020\|EPI_ISL_429028\|2020-04 | ···················· | ···················· | ···················· | ···················· | ···················· | ···················· |
| A | 2007 | hCoV-19/USA/CA-CZB067/2020\|EPI_ISL_429029\|2020-03- | ···················· | ···················· | ···················· | ···················· | ···················· | ···················· |
| A | 2008 | hCoV-19/USA/CA-CZB089/2020\|EPI_ISL_429050\|2020-03- | ···················· | ···················· | ···················· | ···················· | ···················· | ···················· |
| A | 2009 | hCoV-19/USA/CA-CZB0741/2020\|EPI_ISL_429051\|2020-04 | ···················· | ···················· | ···················· | ···················· | ···················· | ···················· |
| A | 2010 | hCoV-19/USA/CA-CZB0714/2020\|EPI_ISL_429052\|2020-03 | ···················· | ···················· | ···················· | ···················· | ···················· | ···················· |
| A | 2011 | hCoV-19/USA/CA-CZB0167/2020\|EPI_ISL_429053\|2020-04 | ···················· | ···················· | ···················· | ···················· | ···················· | ···················· |
| A | 2012 | hCoV-19/USA/CA-CZB0141/2020\|EPI_ISL_429054\|2020-04 | ···················· | ···················· | ···················· | ···················· | ···················· | ···················· |

| | | | | | | | | |
|---|---|---|---|---|---|---|---|---|
| A | 2013 | hCoV-19/USA/CA-CZB0726/2020\|EPI_ISL_429055\|2020-03 | .................... | .................... | ..................... | .................... | .................... | .................... |
| A | 2014 | hCoV-19/USA/CA-CZB0101/2020\|EPI_ISL_429056\|2020-03 | .................... | .................... | ..................... | .................... | .................... | .................... |
| A | 2015 | hCoV-19/USA/CA-CZB076/2020\|EPI_ISL_429057\|2020-03- | .................... | .................... | ..................... | .................... | .................... | .................... |
| A | 2016 | hCoV-19/USA/CA-CZB0114/2020\|EPI_ISL_429058\|2020-03 | .................... | .................... | ..................... | .................... | .................... | .................... |
| A | 2017 | hCoV-19/Jordan/SR-042/2020\|EPI_ISL_430000\|2020-03- | .................... | .................... | ..................... | .................... | .................... | .................... |
| A | 2018 | hCoV-19/Jordan/SR-043/2020\|EPI_ISL_430001\|2020-03- | .................... | .................... | ..................... | .................... | .................... | .................... |
| A | 2019 | hCoV-19/Jordan/SR-044/2020\|EPI_ISL_430002\|2020-03- | .................... | .................... | ..................... | .................... | .................... | .................... |
| A | 2020 | hCoV-19/Jordan/SR-045/2020\|EPI_ISL_430003\|2020-03- | .................... | .................... | ..................... | .................... | .................... | .................... |
| A | 2021 | hCoV-19/Jordan/SR-046/2020\|EPI_ISL_430004\|2020-03- | .................... | .................... | ..................... | .................... | .................... | .................... |
| A | 2022 | hCoV-19/Jordan/SR-048/2020\|EPI_ISL_430006\|2020-03- | .................... | .................... | ..................... | .................... | .................... | .................... |
| A | 2023 | hCoV-19/Jordan/SR-049/2020\|EPI_ISL_430007\|2020-03- | .................... | .................... | ..................... | .................... | .................... | .................... |
| A | 2024 | hCoV-19/Jordan/SR-052/2020\|EPI_ISL_430009\|2020-03- | .................... | .................... | ..................... | .................... | .................... | .................... |
| A | 2025 | hCoV-19/Jordan/SR-053/2020\|EPI_ISL_430011\|2020-03- | .................... | .................... | ..................... | .................... | .................... | .................... |
| A | 2026 | hCoV-19/Jordan/SR-054/2020\|EPI_ISL_430012\|2020-03- | .................... | .................... | ..................... | .................... | .................... | .................... |
| A | 2027 | hCoV-19/Jordan/SR-057/2020\|EPI_ISL_430015\|2020-03- | .................... | .............A...... | ..................... | .................... | .................... | .................... |
| A | 2028 | hCoV-19/USA/CA-SR016/2020\|EPI_ISL_430016\|2020-03-2 | .................... | .................... | .......A............. | .................... | .................... | .................... |
| A | 2029 | hCoV-19/USA/UT-098/2020\|EPI_ISL_430036\|2020-03-14 | .................... | .................... | ..................... | .................... | .................... | .................... |
| A | 2030 | hCoV-19/USA/UT-099/2020\|EPI_ISL_430037\|2020-03-14 | .................... | .................... | ..................... | .................... | .................... | .................... |
| A | 2031 | hCoV-19/USA/UT-0100/2020\|EPI_ISL_430038\|2020-03-15 | .................... | .................... | ..................... | .................... | .................... | .................... |
| A | 2032 | hCoV-19/USA/UT-0370/2020\|EPI_ISL_430039\|2020-03-30 | .................... | .................... | ..................... | .................... | .................... | .................... |
| A | 2033 | hCoV-19/USA/UT-0443/2020\|EPI_ISL_430041\|2020-03-30 | .................... | .................... | ..................... | .................... | .................... | .................... |
| A | 2034 | hCoV-19/USA/UT-0444/2020\|EPI_ISL_430042\|2020-03-30 | .................... | .................... | ..................... | .................... | .................... | .................... |
| A | 2035 | hCoV-19/USA/UT-0445/2020\|EPI_ISL_430043\|2020-03-30 | .................... | .................... | ..................... | .................... | .................... | .................... |
| A | 2036 | hCoV-19/USA/UT-0446/2020\|EPI_ISL_430044\|2020-03-30 | .................... | .................... | ..................... | .................... | .................... | .................... |
| A | 2037 | hCoV-19/USA/UT-0447/2020\|EPI_ISL_430045\|2020-03-31 | .................... | .................... | ..................... | .................... | .................... | .................... |
| A | 2038 | hCoV-19/USA/UT-0491/2020\|EPI_ISL_430046\|2020-04-02 | .................... | .................... | ..................... | .................... | .................... | .................... |
| A | 2039 | hCoV-19/USA/UT-0492/2020\|EPI_ISL_430047\|2020-04-02 | .................... | .................... | ..................... | .................... | .................... | .................... |
| A | 2040 | hCoV-19/USA/UT-0495/2020\|EPI_ISL_430048\|2020-04-02 | .................... | .................... | ..................... | .................... | .................... | .................... |
| A | 2041 | hCoV-19/USA/UT-0500/2020\|EPI_ISL_430050\|2020-03-31 | .................... | .................... | ..................... | .................... | .................... | .................... |
| A | 2042 | hCoV-19/USA/UT-0501/2020\|EPI_ISL_430051\|2020-04-02 | .................... | .................... | ..................... | .................... | .................... | .................... |
| A | 2043 | hCoV-19/USA/UT-0502/2020\|EPI_ISL_430052\|2020-04-03 | .................... | .................... | ..................... | .................... | .................... | .................... |
| A | 2044 | hCoV-19/USA/UT-0503/2020\|EPI_ISL_430053\|2020-04-02 | .................... | .................... | ..................... | .................... | .................... | .................... |
| A | 2045 | hCoV-19/USA/UT-0506/2020\|EPI_ISL_430054\|2020-04-02 | .................... | .................... | ..................... | .................... | .................... | .................... |
| A | 2046 | hCoV-19/USA/UT-0507/2020\|EPI_ISL_430055\|2020-04-02 | .................... | .................... | ..................... | .................... | .................... | .................... |
| A | 2048 | hCoV-19/USA/UT-0510/2020\|EPI_ISL_430057\|2020-04-03 | .................... | .................... | ..................... | .................... | .................... | .................... |
| A | 2049 | hCoV-19/Australia/NSW03/2020\|EPI_ISL_408977\|2020-0 | .................... | .................... | ..................... | .................... | .................... | .................... |
| A | 2050 | hCoV-19/USA/WA4-UW2/2020\|EPI_ISL_413455\|2020-02-28 | .................... | .................... | ..................... | .................... | .................... | .................... |
| A | 2051 | hCoV-19/USA/WA-S2/2020\|EPI_ISL_413456\|2020-02-21 | .................... | .................... | ..................... | .................... | .................... | .................... |
| A | 2052 | hCoV-19/USA/WA6-UW3/2020\|EPI_ISL_413457\|2020-02-29 | .................... | .................... | ..................... | .................... | .................... | .................... |
| A | 2053 | hCoV-19/USA/WA7-UW4/2020\|EPI_ISL_413458\|2020-03-01 | .................... | .................... | ..................... | .................... | .................... | .................... |
| A | 2054 | hCoV-19/Japan/TK-20-31-3/2020\|EPI_ISL_413459\|2020- | .................... | .................... | ..................... | .................... | .................... | .................... |
| A | 2055 | hCoV-19/Belgium/ULG-3000/2020\|EPI_ISL_417004\|2020- | .................... | .................... | ..................... | .................... | .................... | .................... |
| A | 2057 | hCoV-19/Spain/GA-ISCIII-201663/2020\|EPI_ISL_417007 | .................... | .................... | ..................... | .................... | .................... | .................... |
| A | 2058 | hCoV-19/Belgium/ULG-3662/2020\|EPI_ISL_417008\|2020- | .................... | .................... | ..................... | .................... | .................... | NN.................. |
| A | 2059 | hCoV-19/Belgium/ULG-3665/2020\|EPI_ISL_417009\|2020- | .................... | .................... | ..................... | .................... | .................... | .................... |
| A | 2060 | hCoV-19/USA/WA-S17/2020\|EPI_ISL_417070\|2020-03-03 | .................... | .................... | ..................... | .................... | .................... | .................... |
| A | 2061 | hCoV-19/USA/WA-S18/2020\|EPI_ISL_417071\|2020-03-03 | .................... | .................... | ..................... | .................... | .................... | .................... |
| A | 2062 | hCoV-19/USA/WA-S19/2020\|EPI_ISL_417072\|2020-03-02 | .................... | .................... | ..................... | .................... | .................... | .................... |
| A | 2063 | hCoV-19/USA/WA-S20/2020\|EPI_ISL_417073\|2020-03-02 | .................... | .................... | ..................... | .................... | .................... | .................... |
| A | 2064 | hCoV-19/USA/WA-S21/2020\|EPI_ISL_417074\|2020-03-02 | .................... | .................... | ..................... | .................... | .................... | .................... |
| A | 2065 | hCoV-19/USA/WA-S22/2020\|EPI_ISL_417075\|2020-03-02 | .................... | .................... | ..................... | .................... | .................... | .................... |
| A | 2066 | hCoV-19/USA/WA-S23/2020\|EPI_ISL_417076\|2020-03-02 | .................... | .................... | ..................... | .................... | .................... | .................... |
| A | 2067 | hCoV-19/USA/WA-S24/2020\|EPI_ISL_417077\|2020-03-02 | .................... | .................... | ..................... | .................... | .................... | .................... |
| A | 2068 | hCoV-19/USA/WA-S26/2020\|EPI_ISL_417079\|2020-03-02 | .................... | .................... | ..................... | .................... | .................... | .................... |
| A | 2069 | hCoV-19/USA/WA-S28/2020\|EPI_ISL_417081\|2020-03-02 | .................... | .................... | ..................... | .................... | .................... | .................... |
| A | 2070 | hCoV-19/USA/WA-S29/2020\|EPI_ISL_417082\|2020-03-02 | .................... | .................... | ..................... | .................... | .................... | .................... |
| A | 2071 | hCoV-19/England/SHEF-BFE51/2020\|EPI_ISL_418300\|202 | .................... | .................... | ..................... | .................... | .................... | .................... |
| A | 2072 | hCoV-19/England/SHEF-BFE60/2020\|EPI_ISL_418301\|202 | .................... | .................... | ..................... | .................... | .................... | .................... |
| A | 2073 | hCoV-19/England/SHEF-BFE9D/2020\|EPI_ISL_418302\|202 | .................... | .................... | ..................... | .................... | .................... | .................... |
| A | 2074 | hCoV-19/England/SHEF-BFEAC/2020\|EPI_ISL_418303\|202 | .................... | .................... | ..................... | .................... | .................... | .................... |
| A | 2075 | hCoV-19/England/SHEF-BFEBB/2020\|EPI_ISL_418304\|202 | .................... | .................... | ..................... | .................... | .................... | .................... |
| A | 2076 | hCoV-19/England/SHEF-BFED9/2020\|EPI_ISL_418305\|202 | .................... | .................... | ..................... | .................... | .................... | .................... |
| A | 2077 | hCoV-19/England/SHEF-BFEE8/2020\|EPI_ISL_418306\|202 | .................... | .................... | ..................... | .................... | .................... | .................... |
| A | 2078 | hCoV-19/England/SHEF-BFF03/2020\|EPI_ISL_418307\|202 | .................... | .................... | ..................... | .................... | .................... | .................... |
| A | 2079 | hCoV-19/England/SHEF-BFF12/2020\|EPI_ISL_418308\|202 | .................... | .................... | ..................... | .................... | .................... | .................... |
| A | 2080 | hCoV-19/England/SHEF-BFF21/2020\|EPI_ISL_418309\|202 | .................... | .................... | ..................... | .................... | .................... | .................... |
| A | 2081 | hCoV-19/England/SHEF-BFF30/2020\|EPI_ISL_418310\|202 | .................... | .................... | ..................... | .................... | .................... | .................... |
| A | 2082 | hCoV-19/England/SHEF-BFF4F/2020\|EPI_ISL_418311\|202 | .................... | .................... | ..................... | .................... | .................... | .................... |
| A | 2083 | hCoV-19/England/SHEF-BFF5E/2020\|EPI_ISL_418312\|202 | .................... | .................... | ..................... | .................... | .................... | .................... |
| A | 2084 | hCoV-19/England/SHEF-BFF6D/2020\|EPI_ISL_418313\|202 | .................... | .................... | ..................... | .................... | .................... | .................... |
| A | 2085 | hCoV-19/England/SHEF-BFF7C/2020\|EPI_ISL_418314\|202 | .................... | .................... | ..................... | .................... | .................... | .................... |
| A | 2086 | hCoV-19/England/SHEF-BFF8B/2020\|EPI_ISL_418315\|202 | .................... | .................... | ..................... | .................... | .................... | .................... |
| A | 2087 | hCoV-19/England/SHEF-BFF9A/2020\|EPI_ISL_418316\|202 | .................... | .................... | ..................... | .................... | .................... | .................... |
| A | 2088 | hCoV-19/England/SHEF-BFFA9/2020\|EPI_ISL_418317\|202 | .................... | .................... | ..................... | .................... | .................... | .................... |
| A | 2089 | hCoV-19/England/SHEF-BFFB8/2020\|EPI_ISL_418318\|202 | .................... | .................... | ..................... | .................... | .................... | .................... |
| A | 2090 | hCoV-19/England/SHEF-BFFC7/2020\|EPI_ISL_418319\|202 | .................... | .................... | ..................... | .................... | .................... | .................... |
| A | 2091 | hCoV-19/England/SHEF-BFFD6/2020\|EPI_ISL_418320\|202 | .................... | .................... | ..................... | .................... | .................... | .................... |
| A | 2092 | hCoV-19/England/SHEF-BFFE5/2020\|EPI_ISL_418321\|202 | .................... | .................... | ..................... | .................... | .................... | .................... |

| | | | | | | | |
|---|---|---|---|---|---|---|---|
| A | 2093 hCoV-19/Canada/ON-PHL-3917/2020\|EPI_ISL_418322\|202 | .................... | .................... | .................... | .................... | .................... | .................... |
| A | 2094 hCoV-19/Canada/ON-PHL-3919/2020\|EPI_ISL_418323\|202 | .................... | .................... | .................... | .................... | .................... | .................... |
| A | 2095 hCoV-19/Canada/ON-PHL-3318/2020\|EPI_ISL_418324\|202 | .................... | .................... | .................... | .................... | .................... | .................... |
| A | 2096 hCoV-19/Canada/ON-PHL-5472/2020\|EPI_ISL_418325\|202 | .................... | .................... | .................... | .................... | .................... | .................... |
| A | 2097 hCoV-19/Canada/ON-PHL-7972/2020\|EPI_ISL_418326\|202 | .................... | .................... | .................... | .................... | .................... | .................... |
| A | 2098 hCoV-19/Canada/ON-PHL-4181/2020\|EPI_ISL_418327\|202 | .................... | .A.................. | .................... | .................... | .................... | .................... |
| A | 2099 hCoV-19/Canada/ON-PHL-3670/2020\|EPI_ISL_418328\|202 | .................... | .................... | .................... | .................... | .................... | .................... |
| A | 2100 hCoV-19/Canada/ON-PHL-5694/2020\|EPI_ISL_418329\|202 | .................... | .................... | .................... | .................... | .................... | .................... |
| A | 2101 hCoV-19/Canada/ON-PHL-8580/2020\|EPI_ISL_418330\|202 | .................... | .................... | .................... | .................... | .................... | .................... |
| A | 2102 hCoV-19/Canada/ON-PHL-6883/2020\|EPI_ISL_418331\|202 | .................... | .................... | .................... | .................... | .................... | .................... |
| A | 2103 hCoV-19/Canada/ON-PHL-0052/2020\|EPI_ISL_418332\|202 | .................... | .................... | .................... | .................... | .................... | .................... |
| A | 2104 hCoV-19/Canada/ON-PHL-3501/2020\|EPI_ISL_418333\|202 | .................... | .................... | .................... | .................... | .................... | .................... |
| A | 2105 hCoV-19/Canada/ON-PHL-4069/2020\|EPI_ISL_418334\|202 | .................... | .................... | .................... | .................... | .................... | .................... |
| A | 2106 hCoV-19/Canada/ON-PHL-4464/2020\|EPI_ISL_418335\|202 | .................... | .................... | .................... | .................... | .................... | .................... |
| A | 2107 hCoV-19/Canada/ON-PHL-5705/2020\|EPI_ISL_418336\|202 | .................... | .................... | .................... | .................... | .................... | .................... |
| A | 2108 hCoV-19/Canada/ON-PHL-3802/2020\|EPI_ISL_418337\|202 | .................... | .................... | .................... | .................... | .................... | .................... |
| A | 2109 hCoV-19/Canada/ON-PHL-5710/2020\|EPI_ISL_418338\|202 | .................... | .................... | .................... | .................... | .................... | .................... |
| A | 2110 hCoV-19/Canada/ON-PHL-3877/2020\|EPI_ISL_418339\|202 | .................... | .................... | .................... | .................... | .................... | .................... |
| A | 2111 hCoV-19/Luxembourg/LNS9080444/2020\|EPI_ISL_419600\| | .................... | .................... | .................... | .................... | .................... | .................... |
| A | 2112 hCoV-19/Luxembourg/LNS9086704/2020\|EPI_ISL_419601\| | .................... | .................... | .................... | .................... | .................... | .................... |
| A | 2113 hCoV-19/Luxembourg/LNS9324837/2020\|EPI_ISL_419602\| | .................... | .................... | .................... | .................... | .................... | .................... |
| A | 2114 hCoV-19/Luxembourg/LNS9512434/2020\|EPI_ISL_419603\| | .................... | .................... | .................... | .................... | .................... | .................... |
| A | 2115 hCoV-19/Luxembourg/LNS9627078/2020\|EPI_ISL_419604\| | .................... | .................... | .................... | .................... | .................... | .................... |
| A | 2116 hCoV-19/Luxembourg/LNS9862689/2020\|EPI_ISL_419606\| | .................... | .................... | .................... | .................... | .................... | .................... |
| A | 2117 hCoV-19/Luxembourg/LNS9982497/2020\|EPI_ISL_419607\| | .................... | .................... | .................... | .................... | .................... | .................... |
| A | 2118 hCoV-19/USA/WI-GMF-00466/2020\|EPI_ISL_419651\|2020- | .................... | .................... | .................... | .................... | .................... | .................... |
| A | 2119 hCoV-19/USA/WI-GMF-00281/2020\|EPI_ISL_419652\|2020- | .................... | .................... | .................... | .................... | .................... | .................... |
| A | 2120 hCoV-19/Austria/CeMM0001/2020\|EPI_ISL_419654\|2020- | .................... | .................... | .................... | .................... | .................... | .................... |
| A | 2121 hCoV-19/Austria/CeMM0002/2020\|EPI_ISL_419655\|2020- | .................... | .................... | .................... | .................... | .................... | .................... |
| A | 2122 hCoV-19/Austria/CeMM0003/2020\|EPI_ISL_419656\|2020- | .................... | .................... | .................... | .................... | .................... | .................... |
| A | 2123 hCoV-19/Austria/CeMM0004/2020\|EPI_ISL_419657\|2020- | .................... | .................... | .................... | .................... | .................... | .................... |
| A | 2124 hCoV-19/Austria/CeMM0005/2020\|EPI_ISL_419658\|2020- | .................... | .................... | .................... | .................... | .................... | .................... |
| A | 2125 hCoV-19/Austria/CeMM0006/2020\|EPI_ISL_419659\|2020- | .................... | .................... | .................... | .................... | .................... | .................... |
| A | 2126 hCoV-19/Austria/CeMM0007/2020\|EPI_ISL_419660\|2020- | .................... | .................... | .................... | .................... | .................... | .................... |
| A | 2127 hCoV-19/Austria/CeMM0008/2020\|EPI_ISL_419661\|2020- | .................... | .................... | .................... | .................... | .........K.......... | .................... |
| A | 2128 hCoV-19/Austria/CeMM0009/2020\|EPI_ISL_419662\|2020- | .................... | .................... | .................... | .................... | .................... | .................... |
| A | 2129 hCoV-19/Austria/CeMM0011/2020\|EPI_ISL_419664\|2020- | .................... | .................... | .................... | .................... | .................... | .................... |
| A | 2130 hCoV-19/Austria/CeMM0012/2020\|EPI_ISL_419665\|2020- | .................... | .................... | .................... | .................... | .................... | .................... |
| A | 2131 hCoV-19/Austria/CeMM0013/2020\|EPI_ISL_419666\|2020- | .................... | .................... | .................... | .................... | .................... | .................... |
| A | 2132 hCoV-19/Austria/CeMM0014/2020\|EPI_ISL_419667\|2020- | .................... | .................... | .................... | .................... | .................... | .................... |
| A | 2133 hCoV-19/Austria/CeMM0016/2020\|EPI_ISL_419669\|2020- | .................... | .................... | .................... | .................... | N................... | .................... |
| A | 2134 hCoV-19/Argentina/C121/2020\|EPI_ISL_420600\|2020-03 | .................... | .................... | .................... | .................... | .................... | .................... |
| A | 2135 hCoV-19/France/ARA-12371/2020\|EPI_ISL_420604\|2020- | .................... | .................... | .................... | .................... | .................... | .................... |
| A | 2136 hCoV-19/France/ARA-12384/2020\|EPI_ISL_420605\|2020- | .................... | .................... | .................... | .................... | .................... | .................... |
| A | 2137 hCoV-19/France/ARA-12388/2020\|EPI_ISL_420606\|2020- | .................... | .................... | .................... | .................... | .................... | .................... |
| A | 2138 hCoV-19/France/ARA-12485/2020\|EPI_ISL_420607\|2020- | .................... | .................... | .................... | .................... | .................... | .................... |
| A | 2139 hCoV-19/France/ARA-12499/2020\|EPI_ISL_420608\|2020- | .................... | .................... | .................... | .................... | .................... | .................... |
| A | 2140 hCoV-19/France/ARA-12524/2020\|EPI_ISL_420609\|2020- | .................... | .................... | .................... | .................... | .................... | .................... |
| A | 2141 hCoV-19/France/ARA-12558/2020\|EPI_ISL_420610\|2020- | .................... | .................... | .................... | .................... | .................... | .................... |
| A | 2142 hCoV-19/France/ARA-12576/2020\|EPI_ISL_420611\|2020- | .................... | .................... | .................... | .................... | .................... | .................... |
| A | 2143 hCoV-19/France/ARA-12630/2020\|EPI_ISL_420613\|2020- | .................... | .................... | .................... | .................... | .................... | .................... |
| A | 2144 hCoV-19/France/ARA-12632/2020\|EPI_ISL_420614\|2020- | .................... | .................... | .................... | .................... | .................... | .................... |
| A | 2145 hCoV-19/France/ARA-12759/2020\|EPI_ISL_420615\|2020- | .................... | .................... | .................... | .................... | .................... | .................... |
| A | 2146 hCoV-19/France/ARA-1284/2020\|EPI_ISL_420616\|2020-0 | .................... | .................... | .................... | .................... | .................... | .................... |
| A | 2147 hCoV-19/France/ARA-12877/2020\|EPI_ISL_420617\|2020- | .................... | .................... | .................... | .................... | .................... | .................... |
| A | 2148 hCoV-19/France/ARA-12915/2020\|EPI_ISL_420618\|2020- | .................... | .................... | .................... | .................... | .................... | .................... |
| A | 2149 hCoV-19/France/ARA-12973/2020\|EPI_ISL_420619\|2020- | .................... | .................... | .................... | .................... | .................... | .................... |
| A | 2150 hCoV-19/France/ARA-12996/2020\|EPI_ISL_420620\|2020- | .................... | .................... | .................... | .................... | .................... | .................... |
| A | 2151 hCoV-19/France/ARA-1307/2020\|EPI_ISL_420621\|2020-0 | .................... | .................... | .................... | .................... | .................... | .................... |
| A | 2153 hCoV-19/France/ARA-13095/2020\|EPI_ISL_420623\|2020- | .................... | .................... | .................... | .................... | .................... | .................... |
| A | 2154 hCoV-19/France/ARA-13160/2020\|EPI_ISL_420624\|2020- | .................... | .................... | .................... | .................... | .................... | .................... |
| A | 2155 hCoV-19/France/ARA-1322/2020\|EPI_ISL_420625\|2020-0 | .................... | .................... | .................... | .................... | .................... | .................... |
| A | 2156 hCoV-19/USA/VA-DCLS-0024/2020\|EPI_ISL_420628\|2020- | .................... | .................... | .................... | .................... | .................... | .................... |
| A | 2157 hCoV-19/USA/VA-DCLS-0034/2020\|EPI_ISL_420629\|2020- | .................... | .................... | .................... | .................... | .................... | .................... |
| A | 2158 hCoV-19/USA/VA-DCLS-0038/2020\|EPI_ISL_420630\|2020- | .................... | .................... | .................... | .................... | .................... | .................... |
| A | 2159 hCoV-19/England/20130055502/2020\|EPI_ISL_420633\|20 | ...T................ | .................... | .................... | .................... | .................... | .................... |
| A | 2160 hCoV-19/England/20136085404/2020\|EPI_ISL_420636\|20 | .................... | .................... | .................... | .................... | .................... | .................... |
| A | 2161 hCoV-19/England/20136088602/2020\|EPI_ISL_420640\|20 | .................... | .................... | .................... | .................... | .................... | .................... |
| A | 2162 hCoV-19/England/20136089402/2020\|EPI_ISL_420642\|20 | .................... | .................... | .................... | .................... | .................... | .................... |
| A | 2163 hCoV-19/England/20136094704/2020\|EPI_ISL_420643\|20 | .................... | .................... | .................... | .................... | .................... | .................... |
| A | 2164 hCoV-19/England/20136095604/2020\|EPI_ISL_420645\|20 | .................... | .................... | .................... | .................... | .................... | .................... |
| A | 2165 hCoV-19/England/20136096504/2020\|EPI_ISL_420648\|20 | .................... | .................... | .................... | .................... | .................... | .................... |
| A | 2166 hCoV-19/England/20136097204/2020\|EPI_ISL_420652\|20 | .................... | .................... | .................... | .................... | .................... | .................... |
| A | 2167 hCoV-19/England/20139027004/2020\|EPI_ISL_421900\|20 | .................... | .................... | .................... | .................... | .................... | .................... |
| A | 2169 hCoV-19/England/20139027404/2020\|EPI_ISL_421904\|20 | .................... | .................... | .................... | .................... | .................... | .................... |
| A | 2170 hCoV-19/England/20139027604/2020\|EPI_ISL_421905\|20 | .................... | .................... | .................... | .................... | .................... | .................... |
| A | 2171 hCoV-19/England/20139027704/2020\|EPI_ISL_421906\|20 | .................... | .................... | .................... | .................... | .................... | .................... |
| A | 2172 hCoV-19/England/20139027904/2020\|EPI_ISL_421907\|20 | .................... | .................... | .................... | .................... | .................... | .................... |

| | | | | | | | |
|---|---|---|---|---|---|---|---|
| A | 2173 hCoV-19/England/20139029104/2020\|EPI_ISL_421909\|20 | ···················· | ···················· | ···················· | ···················· | ···················· | ···················· |
| A | 2174 hCoV-19/England/20139029304/2020\|EPI_ISL_421910\|20 | ···················· | ···················· | ···················· | ···················· | ···················· | ···················· |
| A | 2175 hCoV-19/England/20139029404/2020\|EPI_ISL_421911\|20 | ···················· | ···················· | ···················· | ···················· | ···················· | ···················· |
| A | 2176 hCoV-19/England/20139030404/2020\|EPI_ISL_421912\|20 | ···················· | ···················· | ···················· | ···················· | ···················· | ···················· |
| A | 2177 hCoV-19/England/20139030704/2020\|EPI_ISL_421913\|20 | ···················· | ·············A······ | ···················· | ···················· | ···················· | ···················· |
| A | 2178 hCoV-19/England/20139031404/2020\|EPI_ISL_421914\|20 | ···················· | ···················· | ···················· | ···················· | ···················· | ···················· |
| A | 2179 hCoV-19/England/20139031504/2020\|EPI_ISL_421915\|20 | ···················· | ···················· | ···················· | ···················· | ···················· | ···················· |
| A | 2180 hCoV-19/England/20139031804/2020\|EPI_ISL_421916\|20 | ···················· | ···················· | ···················· | ···················· | ···················· | ···················· |
| A | 2181 hCoV-19/England/20139032404/2020\|EPI_ISL_421918\|20 | ···················· | ···················· | ···················· | ···················· | ···················· | ···················· |
| A | 2182 hCoV-19/England/20139033004/2020\|EPI_ISL_421919\|20 | ···················· | ···················· | ···················· | ···················· | ···················· | ···················· |
| A | 2184 hCoV-19/England/20139033404/2020\|EPI_ISL_421921\|20 | ···················· | ···················· | ···················· | ···················· | ···················· | ···················· |
| A | 2185 hCoV-19/England/20139036504/2020\|EPI_ISL_421923\|20 | ···················· | ···················· | ···················· | ···················· | ···················· | ···················· |
| A | 2188 hCoV-19/England/20144012204/2020\|EPI_ISL_421930\|20 | ···················· | ···················· | ···················· | ···················· | ··N················· | ···················· |
| A | 2189 hCoV-19/England/20144013504/2020\|EPI_ISL_421932\|20 | ···················· | ···················· | ···················· | ···················· | ···················· | ···················· |
| A | 2190 hCoV-19/England/20144014004/2020\|EPI_ISL_421934\|20 | ···················· | ···················· | ···················· | ···················· | ···················· | ···················· |
| A | 2191 hCoV-19/England/20144014604/2020\|EPI_ISL_421935\|20 | ···················· | ···················· | ···················· | ···················· | ···················· | ···················· |
| A | 2193 hCoV-19/England/20144016604/2020\|EPI_ISL_421937\|20 | ···················· | ···················· | ···················· | ···················· | ···················· | ···················· |
| A | 2194 hCoV-19/England/20144018204/2020\|EPI_ISL_421938\|20 | ···················· | ···················· | ···················· | ···················· | ···················· | ···················· |
| A | 2195 hCoV-19/England/20144018704/2020\|EPI_ISL_421939\|20 | ···················· | ···················· | ···················· | ···················· | ···················· | ···················· |
| A | 2196 hCoV-19/England/20144019304/2020\|EPI_ISL_421940\|20 | ···················· | ···················· | ···················· | ···················· | ···················· | ···················· |
| A | 2197 hCoV-19/England/20144020004/2020\|EPI_ISL_421942\|20 | ···················· | ···················· | ···················· | ···················· | ···················· | ···················· |
| A | 2199 hCoV-19/England/20144037304/2020\|EPI_ISL_421946\|20 | ···················· | ···················· | ···················· | ···················· | ···················· | ···················· |
| A | 2201 hCoV-19/England/20144049404/2020\|EPI_ISL_421948\|20 | ···················· | ···················· | ···················· | ···················· | ···················· | ···················· |
| A | 2202 hCoV-19/England/20144049704/2020\|EPI_ISL_421951\|20 | ···················· | ···················· | ···················· | ···················· | ···················· | ···················· |
| A | 2203 hCoV-19/England/20144052104/2020\|EPI_ISL_421953\|20 | ···················· | ···················· | ···················· | ···················· | ···················· | ···················· |
| A | 2205 hCoV-19/England/20144067504/2020\|EPI_ISL_421957\|20 | ···················· | ···················· | ···················· | ···················· | ···················· | ···················· |
| A | 2206 hCoV-19/England/20144071004/2020\|EPI_ISL_421958\|20 | ···················· | ···················· | ···················· | ···················· | ···················· | ···················· |
| A | 2209 hCoV-19/England/20144076304/2020\|EPI_ISL_421964\|20 | ···················· | ···················· | ···················· | ···················· | ···················· | ···················· |
| A | 2210 hCoV-19/England/20146008104/2020\|EPI_ISL_421974\|20 | ···················· | ···················· | ···················· | ···················· | ···················· | ···················· |
| A | 2212 hCoV-19/England/20146019004/2020\|EPI_ISL_421981\|20 | ···················· | ···················· | ···················· | ···················· | ···················· | ···················· |
| A | 2213 hCoV-19/England/20146019704/2020\|EPI_ISL_421982\|20 | ···················· | ···················· | ···················· | ···················· | ···················· | ···················· |
| A | 2214 hCoV-19/England/20129125504/2020\|EPI_ISL_424100\|20 | ···················· | ···················· | ···················· | ···················· | ···················· | ···················· |
| A | 2215 hCoV-19/England/20129126204/2020\|EPI_ISL_424101\|20 | ···················· | ···················· | ···················· | ···················· | ···················· | R··················· |
| A | 2217 hCoV-19/England/20129147804/2020\|EPI_ISL_424116\|20 | ···················· | ···················· | ···················· | ···················· | ···················· | ···················· |
| A | 2218 hCoV-19/England/20129148904/2020\|EPI_ISL_424117\|20 | ···················· | ···················· | ···················· | ···················· | ···················· | ···················· |
| A | 2219 hCoV-19/USA/WA-UW-1294/2020\|EPI_ISL_424180\|2020-03 | ···················· | ···················· | ···················· | ···················· | ···················· | ···················· |
| A | 2220 hCoV-19/USA/UN-UW-1402/2020\|EPI_ISL_424181\|2020-03 | ···················· | ···················· | ···················· | ···················· | ···················· | ···················· |
| A | 2221 hCoV-19/USA/WA-UW-1478/2020\|EPI_ISL_424182\|2020-03 | ···················· | ···················· | ···················· | ···················· | ···················· | ···················· |
| A | 2222 hCoV-19/USA/WA-UW-1436/2020\|EPI_ISL_424183\|2020-03 | ···················· | ···················· | ···················· | ···················· | ···················· | ···················· |
| A | 2223 hCoV-19/USA/WA-UW-1422/2020\|EPI_ISL_424184\|2020-03 | ···················· | ···················· | ···················· | ···················· | ···················· | ···················· |
| A | 2224 hCoV-19/USA/WA-UW-1644/2020\|EPI_ISL_424186\|2020-03 | ···················· | ···················· | ···················· | ···················· | ···················· | ···················· |
| A | 2225 hCoV-19/USA/WA-UW-1646/2020\|EPI_ISL_424187\|2020-03 | ···················· | ···················· | ···················· | ···················· | ···················· | ···················· |
| A | 2226 hCoV-19/USA/WA-UW-1645/2020\|EPI_ISL_424188\|2020-03 | ···················· | ···················· | ···················· | ···················· | ···················· | ···················· |
| A | 2227 hCoV-19/USA/WA-UW-01/2020\|EPI_ISL_424189\|2020-03 | ···················· | ···················· | ···················· | ···················· | ···················· | ···················· |
| A | 2228 hCoV-19/USA/WA-UW-1683/2020\|EPI_ISL_424191\|2020-03 | ···················· | ···················· | ···················· | ···················· | ···················· | ···················· |
| A | 2229 hCoV-19/USA/WA-UW-1688/2020\|EPI_ISL_424192\|2020-03 | ···················· | ···················· | ···················· | ···················· | ···················· | ···················· |
| A | 2230 hCoV-19/USA/WA-UW-1728/2020\|EPI_ISL_424193\|2020-03 | ···················· | ···················· | ···················· | ···················· | ···················· | ···················· |
| A | 2231 hCoV-19/USA/WA-UW-1661/2020\|EPI_ISL_424194\|2020-03 | ···················· | ···················· | ···················· | ···················· | ···················· | ···················· |
| A | 2232 hCoV-19/USA/WA-UW-1726/2020\|EPI_ISL_424195\|2020-03 | ···················· | ···················· | ···················· | ···················· | ···················· | ···················· |
| A | 2233 hCoV-19/USA/WA-UW-1666/2020\|EPI_ISL_424196\|2020-03 | ···················· | ···················· | ···················· | ···················· | ···················· | ···················· |
| A | 2234 hCoV-19/USA/WA-UW-1640/2020\|EPI_ISL_424197\|2020-03 | ···················· | ···················· | ···················· | ···················· | ···················· | ···················· |
| A | 2235 hCoV-19/USA/WA-UW-1656/2020\|EPI_ISL_424198\|2020-03 | ···················· | ···················· | ···················· | ···················· | ···················· | ···················· |
| A | 2236 hCoV-19/England/CAMB-76D95/2020\|EPI_ISL_425427\|202 | ···················· | ···················· | ···················· | ···················· | ···················· | ···················· |
| A | 2237 hCoV-19/England/CAMB-76DB3/2020\|EPI_ISL_425428\|202 | ···················· | ···················· | ···················· | ···················· | ···················· | ···················· |
| A | 2238 hCoV-19/England/CAMB-76DFF/2020\|EPI_ISL_425430\|202 | ···················· | ···················· | ···················· | ···················· | ···················· | ···················· |
| A | 2239 hCoV-19/England/CAMB-76E1A/2020\|EPI_ISL_425431\|202 | ···················· | ···················· | ···················· | ···················· | ···················· | ···················· |
| A | 2241 hCoV-19/England/CAMB-76E56/2020\|EPI_ISL_425433\|202 | ···················· | ···················· | ···················· | ···················· | ···················· | ···················· |
| A | 2242 hCoV-19/England/CAMB-76E74/2020\|EPI_ISL_425434\|202 | ···················· | ···················· | ···················· | ···················· | ···················· | ···················· |
| A | 2243 hCoV-19/England/CAMB-76E92/2020\|EPI_ISL_425435\|202 | ···················· | ···················· | ···················· | ···················· | ···················· | ···················· |
| A | 2244 hCoV-19/England/CAMB-76EB0/2020\|EPI_ISL_425436\|202 | ···················· | ···················· | ···················· | ···················· | ···················· | ···················· |
| A | 2245 hCoV-19/England/CAMB-76EDE/2020\|EPI_ISL_425437\|202 | ···················· | ···················· | ···················· | ···················· | ···················· | ···················· |
| A | 2246 hCoV-19/England/CAMB-76F17/2020\|EPI_ISL_425439\|202 | ···················· | ···················· | ···················· | ···················· | ···················· | ···················· |
| A | 2247 hCoV-19/England/CAMB-76F35/2020\|EPI_ISL_425440\|202 | ···················· | ···················· | ···················· | ···················· | ···················· | ···················· |
| A | 2248 hCoV-19/England/CAMB-76F62/2020\|EPI_ISL_425441\|202 | ···················· | ···················· | ···················· | ···················· | ···················· | ···················· |
| A | 2249 hCoV-19/England/CAMB-76F9F/2020\|EPI_ISL_425443\|202 | ···················· | ···················· | ···················· | ···················· | ···················· | ···················· |
| A | 2250 hCoV-19/England/CAMB-76FCC/2020\|EPI_ISL_425446\|202 | ···················· | ···················· | ···················· | ···················· | ···················· | ···················· |
| A | 2251 hCoV-19/England/NOTT-10DF14/2020\|EPI_ISL_425469\|20 | ···················· | ···················· | ···················· | ···················· | ···················· | ···················· |
| A | 2253 hCoV-19/Australia/VIC512/2020\|EPI_ISL_426782\|2020- | ···················· | ···················· | ···················· | ···················· | ···················· | ···················· |
| A | 2254 hCoV-19/Australia/VIC513/2020\|EPI_ISL_426783\|2020- | ···················· | ···················· | ···················· | ···················· | ···················· | ···················· |
| A | 2255 hCoV-19/Australia/VIC514/2020\|EPI_ISL_426784\|2020- | ···················· | ···················· | ···················· | ···················· | ···················· | ···················· |
| A | 2256 hCoV-19/Australia/VIC516/2020\|EPI_ISL_426786\|2020- | ···················· | ···················· | ···················· | ···················· | ···················· | ···················· |
| A | 2258 hCoV-19/Australia/VIC520/2020\|EPI_ISL_426789\|2020- | ···················· | ···················· | ···················· | ···················· | ···················· | ···················· |
| A | 2259 hCoV-19/Australia/VIC522/2020\|EPI_ISL_426790\|2020- | ···················· | ···················· | ···················· | ···················· | ···················· | ···················· |
| A | 2261 hCoV-19/Australia/VIC527/2020\|EPI_ISL_426793\|2020- | ···················· | ···················· | ···················· | ···················· | ···················· | ···················· |
| A | 2262 hCoV-19/Australia/VIC530/2020\|EPI_ISL_426796\|2020- | ···················· | ···················· | ···················· | ···················· | ···················· | ···················· |
| A | 2263 hCoV-19/Australia/VIC531/2020\|EPI_ISL_426797\|2020- | ···················· | ···················· | ···················· | ···················· | ···················· | ···················· |
| A | 2264 hCoV-19/Australia/VIC532/2020\|EPI_ISL_426798\|2020- | ···················· | ···················· | ···················· | ···················· | ···················· | ···················· |
| A | 2266 hCoV-19/USA/CA-CZB072/2020\|EPI_ISL_429030\|2020-03- | ···················· | ···················· | ···················· | ···················· | ···················· | ···················· |

| | | | | | | | | |
|---|---|---|---|---|---|---|---|---|
| A | 2267 | hCoV-19/USA/CA-CZB0139/2020\|EPI_ISL_429031\|2020-04 | ···················· | ···················· | ····················· | ···················· | ···················· | ···················· |
| A | 2268 | hCoV-19/USA/CA-CZB082/2020\|EPI_ISL_429032\|2020-03- | ···················· | ···················· | ····················· | ···················· | ···················· | ···················· |
| A | 2269 | hCoV-19/USA/CA-CZB086/2020\|EPI_ISL_429033\|2020-03- | ···················· | ···················· | ····················· | ···················· | ···················· | ···················· |
| A | 2270 | hCoV-19/USA/CA-CZB0706/2020\|EPI_ISL_429034\|2020-03 | ···················· | ···················· | ····················· | ···················· | ···················· | ···················· |
| A | 2271 | hCoV-19/USA/CA-CZB042/2020\|EPI_ISL_429035\|2020-03- | ···················· | ···················· | ····················· | ···················· | ···················· | ···················· |
| A | 2272 | hCoV-19/USA/CA-CZB0107/2020\|EPI_ISL_429036\|2020-03 | ···················· | ···················· | ····················· | ···················· | ···················· | ···················· |
| A | 2273 | hCoV-19/USA/CA-CZB09/2020\|EPI_ISL_429037\|2020-03-2 | ···················· | ···················· | ····················· | ···················· | ···················· | ···················· |
| A | 2274 | hCoV-19/USA/CA-CZB0707/2020\|EPI_ISL_429038\|2020-03 | ···················· | ···················· | ····················· | ···················· | ···················· | ···················· |
| A | 2275 | hCoV-19/USA/CA-CZB036/2020\|EPI_ISL_429039\|2020-03- | ···················· | ···················· | ····················· | ···················· | ···················· | ···················· |
| A | 2276 | hCoV-19/USA/CA-CZB0701/2020\|EPI_ISL_429040\|2020-03 | ···················· | ···················· | ····················· | ···················· | ···················· | ···················· |
| A | 2277 | hCoV-19/USA/CA-CZB096/2020\|EPI_ISL_429041\|2020-03- | ···················· | ···················· | ····················· | ···················· | ···················· | ···················· |
| A | 2278 | hCoV-19/USA/CA-CZB0710/2020\|EPI_ISL_429042\|2020-03 | ···················· | ···················· | ····················· | ···················· | ···················· | ···················· |
| A | 2279 | hCoV-19/USA/CA-CZB0156/2020\|EPI_ISL_429043\|2020-04 | ···················· | ···················· | ····················· | ···················· | ···················· | ···················· |
| A | 2280 | hCoV-19/USA/CA-CZB0158/2020\|EPI_ISL_429044\|2020-04 | ···················· | ···················· | ····················· | ···················· | ···················· | ···················· |
| A | 2281 | hCoV-19/USA/CA-CZB039/2020\|EPI_ISL_429045\|2020-03- | ···················· | ···················· | ····················· | ···················· | ···················· | ···················· |
| A | 2282 | hCoV-19/USA/CA-CZB0125/2020\|EPI_ISL_429046\|2020-04 | ···················· | ···················· | ····················· | ···················· | ···················· | ···················· |
| A | 2283 | hCoV-19/USA/CA-CZB0154/2020\|EPI_ISL_429047\|2020-04 | ···················· | ···················· | ····················· | ···················· | ···················· | ···················· |
| A | 2284 | hCoV-19/USA/CA-CZB043/2020\|EPI_ISL_429048\|2020-03- | ···················· | ···················· | ····················· | ···················· | ···················· | ···················· |
| A | 2285 | hCoV-19/USA/CA-CZB090/2020\|EPI_ISL_429049\|2020-03- | ···················· | ···················· | ····················· | ···················· | ···················· | ···················· |
| A | 2286 | hCoV-19/Jordan/SR-056/2020\|EPI_ISL_430014\|2020-03- | ···················· | ············A······· | ····················· | ···················· | ···················· | ···················· |
| A | 2287 | hCoV-19/USA/UT-066/2020\|EPI_ISL_430019\|2020-03-16 | ···················· | ···················· | ····················· | ···················· | ···················· | ···················· |
| A | 2288 | hCoV-19/USA/UT-069/2020\|EPI_ISL_430021\|2020-03-16 | ···················· | ···················· | ····················· | ···················· | ···················· | ···················· |
| A | 2289 | hCoV-19/USA/UT-070/2020\|EPI_ISL_430022\|2020-03-16 | ···················· | ···················· | ····················· | ···················· | ···················· | ···················· |
| A | 2290 | hCoV-19/USA/UT-071/2020\|EPI_ISL_430023\|2020-03-16 | ···················· | ···················· | ····················· | ···················· | ···················· | ···················· |
| A | 2291 | hCoV-19/USA/UT-072/2020\|EPI_ISL_430024\|2020-03-16 | ···················· | ···················· | ····················· | ···················· | ···················· | ···················· |
| A | 2292 | hCoV-19/USA/UT-074/2020\|EPI_ISL_430025\|2020-03-17 | ···················· | ···················· | ····················· | ···················· | ···················· | ···················· |
| A | 2293 | hCoV-19/USA/UT-075/2020\|EPI_ISL_430026\|2020-03-16 | ···················· | ···················· | ····················· | ···················· | ···················· | ···················· |
| A | 2294 | hCoV-19/USA/UT-077/2020\|EPI_ISL_430028\|2020-03-17 | ···················· | ···················· | ····················· | ···················· | ···················· | ···················· |
| A | 2295 | hCoV-19/USA/UT-078/2020\|EPI_ISL_430029\|2020-03-15 | ···················· | ···················· | ····················· | ···················· | ···················· | ···················· |
| A | 2296 | hCoV-19/USA/UT-080/2020\|EPI_ISL_430031\|2020-03-15 | ···················· | ···················· | ····················· | ···················· | ···················· | ···················· |
| A | 2297 | hCoV-19/USA/UT-083/2020\|EPI_ISL_430032\|2020-03-17 | ···················· | ············A······· | ····················· | ···················· | ···················· | ···················· |
| A | 2298 | hCoV-19/USA/UT-084/2020\|EPI_ISL_430033\|2020-03-17 | ···················· | ···················· | ····················· | ···················· | ···················· | ···················· |
| A | 2300 | hCoV-19/USA/UT-097/2020\|EPI_ISL_430035\|2020-03-15 | ···················· | ···················· | ····················· | ···················· | ···················· | ···················· |
| A | 2301 | hCoV-19/dog/Hong Kong/20-03695/2020\|EPI_ISL_450403 | ···················· | ···················· | ····················· | ···················· | ···················· | ···················· |
| A | 2302 | hCoV-19/Hong Kong/VM20009579/2020\|EPI_ISL_450404\|2 | ···················· | ···················· | ····················· | ···················· | ···················· | ···················· |
| A | 2303 | hCoV-19/Hong Kong/VM2003179/2020\|EPI_ISL_450405\|20 | ···················· | ···················· | ····················· | ···················· | ···················· | ···················· |
| A | 2304 | hCoV-19/Wuhan/YB012504/2020\|EPI_ISL_450489\|2020-01 | ···················· | ···················· | ····················· | ···················· | ···················· | ···················· |
| A | 2305 | hCoV-19/Zhejiang/WZ-01/2020\|EPI_ISL_404227\|2020-01 | ···················· | ···················· | ····················· | ···················· | ···················· | ···················· |
| A | 2306 | hCoV-19/Zhejiang/WZ-02/2020\|EPI_ISL_404228\|2020-01 | ···················· | ···················· | ····················· | ···················· | ···················· | ···················· |
| A | 2307 | hCoV-19/USA/IL1/2020\|EPI_ISL_404253\|2020-01-21 | ···················· | ·R·················· | ····················· | ···················· | ···················· | ···················· |
| A | 2308 | hCoV-19/England/09c/2020\|EPI_ISL_412116\|2020-02-09 | ···················· | ···················· | ····················· | ···················· | ···················· | ···················· |
| A | 2309 | hCoV-19/USA/WA8-UW5/2020\|EPI_ISL_413486\|2020-03-01 | ···················· | ···················· | ····················· | ···················· | ···················· | ···················· |
| A | 2310 | hCoV-19/USA/WA9-UW6/2020\|EPI_ISL_413487\|2020-03-01 | ···················· | ···················· | ····················· | ···················· | ···················· | ···················· |
| A | 2311 | hCoV-19/Germany/NW-HHU-01/2020\|EPI_ISL_413488\|2020 | ···················· | ···················· | ····················· | ···················· | ···················· | ···················· |
| A | 2312 | hCoV-19/Italy/LOM-UniSR-1/2020\|EPI_ISL_413489\|2020 | ···················· | ···················· | ····················· | ···················· | ···················· | ···················· |
| A | 2313 | hCoV-19/New Zealand/20VR0174/2020\|EPI_ISL_413490\|2 | ···················· | ···················· | ····················· | ···················· | ···················· | ···················· |
| A | 2314 | hCoV-19/Luxembourg/LNS5731562/2020\|EPI_ISL_417529\| | ···················· | ···················· | ····················· | ···················· | ···················· | ···················· |
| A | 2315 | hCoV-19/Luxembourg/LNS2128808/2020\|EPI_ISL_417530\| | ···················· | ···················· | ····················· | ···················· | ···················· | ···················· |
| A | 2316 | hCoV-19/Luxembourg/LNS2013896/2020\|EPI_ISL_417531\| | ···················· | ···················· | ····················· | ···················· | ···················· | ···················· |
| A | 2317 | hCoV-19/Luxembourg/LNS8489624/2020\|EPI_ISL_417532\| | ···················· | ···················· | ····················· | ···················· | ···················· | ···················· |
| A | 2318 | hCoV-19/Luxembourg/LNS6282845/2020\|EPI_ISL_417533\| | ···················· | ···················· | ····················· | ···················· | ···················· | ···················· |
| A | 2319 | hCoV-19/Luxembourg/LNS0158952/2020\|EPI_ISL_417534\| | ···················· | ···················· | ····················· | ···················· | ···················· | ···················· |
| A | 2320 | hCoV-19/Iceland/1/2020\|EPI_ISL_417535\|2020-03-13 | ···················· | ···················· | ····················· | ···················· | ···················· | ···················· |
| A | 2321 | hCoV-19/Iceland/10/2020\|EPI_ISL_417536\|2020-03-16 | ···················· | ···················· | ····················· | ···················· | ···················· | ···················· |
| A | 2322 | hCoV-19/Iceland/100/2020\|EPI_ISL_417537\|2020-03-16 | ···················· | ···················· | ····················· | ···················· | ···················· | ···················· |
| A | 2323 | hCoV-19/Iceland/104/2020\|EPI_ISL_417538\|2020-03-17 | ···················· | ···················· | ····················· | ···················· | ···················· | ···················· |
| A | 2324 | hCoV-19/Iceland/11/2020\|EPI_ISL_417539\|2020-03-16 | ···················· | ···················· | ····················· | ···················· | ···················· | ···················· |
| A | 2325 | hCoV-19/Iceland/115/2020\|EPI_ISL_417540\|2020-03-17 | ···················· | ···················· | ····················· | ···················· | ···················· | ···················· |
| A | 2326 | hCoV-19/Iceland/116/2020\|EPI_ISL_417541\|2020-03-17 | ···················· | ···················· | ····················· | ···················· | ···················· | ···················· |
| A | 2327 | hCoV-19/Iceland/117/2020\|EPI_ISL_417542\|2020-03-17 | ···················· | ···················· | ····················· | ···················· | ···················· | ···················· |
| A | 2328 | hCoV-19/Iceland/118/2020\|EPI_ISL_417543\|2020-03-17 | ···················· | ···················· | ····················· | ···················· | ···················· | ···················· |
| A | 2329 | hCoV-19/Iceland/119/2020\|EPI_ISL_417544\|2020-03-17 | ···················· | ···················· | ····················· | ···················· | ···················· | ···················· |
| A | 2330 | hCoV-19/Iceland/120/2020\|EPI_ISL_417545\|2020-03-17 | ···················· | ···················· | ····················· | ···················· | ···················· | ···················· |
| A | 2331 | hCoV-19/Iceland/170/2020\|EPI_ISL_417548\|2020-03-17 | ···················· | ···················· | ····················· | ···················· | ···················· | ···················· |
| A | 2332 | hCoV-19/Iceland/171/2020\|EPI_ISL_417549\|2020-03-14 | ···················· | ···················· | ····················· | ···················· | ···················· | ···················· |
| A | 2333 | hCoV-19/Iceland/232/2020\|EPI_ISL_417560\|2020-03-17 | ···················· | ···················· | ····················· | ···················· | ···················· | ···················· |
| A | 2334 | hCoV-19/Iceland/233/2020\|EPI_ISL_417561\|2020-03-17 | ···················· | ···················· | ····················· | ···················· | ···················· | ···················· |
| A | 2335 | hCoV-19/Iceland/234/2020\|EPI_ISL_417562\|2020-03-17 | ···················· | ···················· | ····················· | ···················· | ···················· | ···················· |
| A | 2336 | hCoV-19/Iceland/235/2020\|EPI_ISL_417563\|2020-03-16 | ···················· | ···················· | ····················· | ···················· | ···················· | ···················· |
| A | 2337 | hCoV-19/Iceland/236/2020\|EPI_ISL_417564\|2020-03-17 | ···················· | ···················· | ····················· | ···················· | ···················· | ···················· |
| A | 2338 | hCoV-19/Iceland/237/2020\|EPI_ISL_417565\|2020-03-17 | ···················· | ···················· | ····················· | ···················· | ···················· | ···················· |
| A | 2339 | hCoV-19/Iceland/238/2020\|EPI_ISL_417566\|2020-03-17 | ···················· | ···················· | ····················· | ···················· | ···················· | ···················· |
| A | 2340 | hCoV-19/Iceland/239/2020\|EPI_ISL_417567\|2020-03-17 | ···················· | ···················· | ····················· | ···················· | ···················· | ···················· |
| A | 2341 | hCoV-19/Iceland/240/2020\|EPI_ISL_417568\|2020-03-17 | ···················· | ···················· | ····················· | ···················· | ···················· | ···················· |
| A | 2342 | hCoV-19/Iceland/241/2020\|EPI_ISL_417569\|2020-03-17 | ···················· | ···················· | ····················· | ···················· | ···················· | ···················· |
| A | 2343 | hCoV-19/Spain/CT-HUVH-VH1133/2020\|EPI_ISL_418860\|2 | ···················· | ···················· | ····················· | ···················· | ···················· | ···················· |
| A | 2344 | hCoV-19/Spain/CT-HUVH-VH2683/2020\|EPI_ISL_418861\|2 | ···················· | ···················· | ····················· | ···················· | ···················· | ···················· |
| A | 2345 | hCoV-19/Belgium/VS-030542/2020\|EPI_ISL_418863\|2020 | ···················· | ···················· | ····················· | ···················· | ···················· | ···················· |

| | | | | | | | |
|---|---|---|---|---|---|---|---|
| A | 2346 hCoV-19/USA/VA-DCLS-0001/2020\|EPI_ISL_418864\|2020- | ···················· | ···················· | ···················· | ···················· | ···················· | ···················· |
| A | 2347 hCoV-19/USA/WA-UW298/2020\|EPI_ISL_418866\|2020-03-1 | ···················· | ···················· | ···················· | ···················· | ···················· | ···················· |
| A | 2348 hCoV-19/USA/WA-UW299/2020\|EPI_ISL_418867\|2020-03-1 | ···················· | ···················· | ···················· | ···················· | ···················· | ···················· |
| A | 2349 hCoV-19/USA/WA-UW300/2020\|EPI_ISL_418868\|2020-03-1 | ···················· | ···················· | ···················· | ···················· | ···················· | ···················· |
| A | 2350 hCoV-19/USA/WA-UW301/2020\|EPI_ISL_418869\|2020-03-1 | ···················· | ···················· | ···················· | ···················· | ···················· | ···················· |
| A | 2351 hCoV-19/USA/WA-UW302/2020\|EPI_ISL_418870\|2020-03-1 | ···················· | ···················· | ···················· | ···················· | ···················· | ···················· |
| A | 2352 hCoV-19/USA/CT-UW303/2020\|EPI_ISL_418871\|2020-03-1 | ···················· | ···················· | ···················· | ···················· | ···················· | ···················· |
| A | 2353 hCoV-19/USA/WA-UW304/2020\|EPI_ISL_418872\|2020-03-1 | ···················· | ···················· | ···················· | ···················· | ···················· | ···················· |
| A | 2354 hCoV-19/USA/WA-UW305/2020\|EPI_ISL_418873\|2020-03-1 | ···················· | ···················· | ···················· | ···················· | ···················· | ···················· |
| A | 2355 hCoV-19/USA/WA-UW307/2020\|EPI_ISL_418875\|2020-03-1 | ···················· | ···················· | ···················· | ···················· | ···················· | ···················· |
| A | 2356 hCoV-19/USA/WA-UW308/2020\|EPI_ISL_418876\|2020-03-1 | ···················· | ···················· | ···················· | ···················· | ···················· | ···················· |
| A | 2357 hCoV-19/USA/WA-UW309/2020\|EPI_ISL_418877\|2020-03-2 | ···················· | ···················· | ···················· | ···················· | ···················· | ···················· |
| A | 2358 hCoV-19/USA/WA-UW310/2020\|EPI_ISL_418878\|2020-03-1 | ···················· | ···················· | ···················· | ···················· | ···················· | ···················· |
| A | 2359 hCoV-19/USA/WA-UW323/2020\|EPI_ISL_418890\|2020-03-1 | ···················· | ···················· | ···················· | ···················· | ···················· | ···················· |
| A | 2360 hCoV-19/USA/WA-UW324/2020\|EPI_ISL_418891\|2020-03-2 | ···················· | ···················· | ···················· | ···················· | ···················· | ···················· |
| A | 2361 hCoV-19/USA/WA-UW325/2020\|EPI_ISL_418892\|2020-03-1 | ···················· | ···················· | ···················· | ···················· | ···················· | ···················· |
| A | 2362 hCoV-19/USA/MN-UW326/2020\|EPI_ISL_418893\|2020-03-1 | ···················· | ···················· | ···················· | ···················· | ···················· | ···················· |
| A | 2363 hCoV-19/USA/CT-UW328/2020\|EPI_ISL_418895\|2020-03-1 | ···················· | ···················· | ···················· | ···················· | ···················· | ···················· |
| A | 2364 hCoV-19/USA/WA-UW330/2020\|EPI_ISL_418897\|2020-03-1 | ···················· | ···················· | ···················· | ···················· | ···················· | ···················· |
| A | 2365 hCoV-19/USA/CT-UW331/2020\|EPI_ISL_418898\|2020-03-1 | ···················· | ···················· | ···················· | ···················· | ···················· | ···················· |
| A | 2366 hCoV-19/USA/WA-UW332/2020\|EPI_ISL_418899\|2020-03-1 | ···················· | ···················· | ···················· | ···················· | ···················· | ···················· |
| A | 2367 hCoV-19/England/20140068404/2020\|EPI_ISL_423400\|20 | ···················· | ···················· | ···················· | ···················· | ···················· | ···················· |
| A | 2368 hCoV-19/England/20140068904/2020\|EPI_ISL_423401\|20 | ···················· | ···················· | ···················· | ···················· | ···················· | ···················· |
| A | 2370 hCoV-19/England/20129159604/2020\|EPI_ISL_424120\|20 | ···················· | ···················· | ···················· | ···················· | ···················· | ···················· |
| A | 2371 hCoV-19/England/20129160704/2020\|EPI_ISL_424121\|20 | ···················· | ···················· | ···················· | ···················· | ···················· | ···················· |
| A | 2372 hCoV-19/England/20129163604/2020\|EPI_ISL_424126\|20 | ···················· | ···················· | ···················· | ···················· | ···················· | ···················· |
| A | 2373 hCoV-19/England/20129170204/2020\|EPI_ISL_424133\|20 | ···················· | ···················· | ···················· | ···················· | ···················· | ···················· |
| A | 2374 hCoV-19/England/20129171604/2020\|EPI_ISL_424140\|20 | ···················· | ···················· | ···················· | ···················· | ···················· | ··N················· |
| A | 2375 hCoV-19/England/20129174804/2020\|EPI_ISL_424152\|20 | ···················· | ···················· | ···················· | ···················· | ···················· | ···················· |
| A | 2376 hCoV-19/USA/WA-UW-1620/2020\|EPI_ISL_424166\|2020-03 | ···················· | ···················· | ···················· | ···················· | ···················· | ···················· |
| A | 2377 hCoV-19/USA/WA-UW-1616/2020\|EPI_ISL_424167\|2020-03 | ···················· | ···················· | ···················· | ···················· | ···················· | ···················· |
| A | 2378 hCoV-19/USA/WA-UW-1538/2020\|EPI_ISL_424169\|2020-03 | ···················· | ···················· | ···················· | ···················· | ···················· | ···················· |
| A | 2379 hCoV-19/USA/WA-UW-1595/2020\|EPI_ISL_424170\|2020-03 | ···················· | ···················· | ···················· | ···················· | ···················· | ···················· |
| A | 2380 hCoV-19/USA/WA-UW-1575/2020\|EPI_ISL_424172\|2020-03 | ···················· | ···················· | ···················· | ···················· | ···················· | ···················· |
| A | 2381 hCoV-19/USA/IL-UW-1311/2020\|EPI_ISL_424173\|2020-03 | ···················· | ···················· | ···················· | ···················· | ···················· | ···················· |
| A | 2382 hCoV-19/USA/CT-UW-1365/2020\|EPI_ISL_424174\|2020-03 | ···················· | ···················· | ···················· | ···················· | ···················· | ···················· |
| A | 2383 hCoV-19/USA/WA-UW-1362/2020\|EPI_ISL_424175\|2020-03 | ···················· | ···················· | ···················· | ···················· | ···················· | ···················· |
| A | 2384 hCoV-19/USA/WA-UW-1472/2020\|EPI_ISL_424176\|2020-03 | ···················· | ···················· | ···················· | ···················· | ···················· | ···················· |
| A | 2385 hCoV-19/USA/WA-UW-1342/2020\|EPI_ISL_424178\|2020-03 | ···················· | ···················· | ···················· | ···················· | ···················· | ···················· |
| A | 2386 hCoV-19/USA/WA-UW-1341/2020\|EPI_ISL_424179\|2020-03 | ···················· | ···················· | ···················· | ···················· | ···················· | ···················· |
| A | 2387 hCoV-19/Sweden/RV-FOI-1/2020\|EPI_ISL_424703\|2020-0 | ···················· | ···················· | ···················· | ···················· | ···················· | ···················· |
| A | 2398 hCoV-19/England/CAMB-76D1D/2020\|EPI_ISL_425423\|202 | ···················· | ···················· | ···················· | ···················· | ···················· | ···················· |
| A | 2399 hCoV-19/England/CAMB-76D3B/2020\|EPI_ISL_425424\|202 | ···················· | ···················· | ···················· | ···················· | ···················· | ···················· |
| A | 2400 hCoV-19/England/CAMB-76D59/2020\|EPI_ISL_425425\|202 | ···················· | ···················· | ···················· | ···················· | ···················· | ···················· |
| A | 2401 hCoV-19/England/CAMB-76FF9/2020\|EPI_ISL_425449\|202 | ···················· | ···················· | ···················· | ···················· | ···················· | ···················· |
| A | 2402 hCoV-19/England/CAMB-77004/2020\|EPI_ISL_425450\|202 | ···················· | ···················· | ···················· | ···················· | ···················· | ···················· |
| A | 2403 hCoV-19/England/CAMB-77013/2020\|EPI_ISL_425451\|202 | ···················· | ··G················· | ···················· | ···················· | ···················· | ···················· |
| A | 2404 hCoV-19/England/CAMB-77022/2020\|EPI_ISL_425452\|202 | ···················· | ···················· | ···················· | ···················· | ···················· | ···················· |
| A | 2405 hCoV-19/England/CAMB-77031/2020\|EPI_ISL_425453\|202 | ···················· | ···················· | ···················· | ···················· | ···················· | ···················· |
| A | 2406 hCoV-19/England/CAMB-77040/2020\|EPI_ISL_425454\|202 | ···················· | ···················· | ···················· | ···················· | ···················· | ···················· |
| A | 2407 hCoV-19/England/CAMB-7705F/2020\|EPI_ISL_425455\|202 | ···················· | ···················· | ···················· | ···················· | ···················· | ···················· |
| A | 2408 hCoV-19/England/CAMB-7707D/2020\|EPI_ISL_425457\|202 | ···················· | ···················· | ···················· | ···················· | ···················· | ···················· |
| A | 2409 hCoV-19/England/CAMB-7708C/2020\|EPI_ISL_425458\|202 | ···················· | ···················· | ···················· | ···················· | ···················· | ···················· |
| A | 2410 hCoV-19/England/CAMB-770B9/2020\|EPI_ISL_425460\|202 | ···················· | ···················· | ···················· | ···················· | ···················· | ···················· |
| A | 2411 hCoV-19/England/NOTT-10DEAE/2020\|EPI_ISL_425462\|20 | ···················· | ···················· | ···················· | ···················· | ···················· | ···················· |
| A | 2412 hCoV-19/England/NOTT-10DECC/2020\|EPI_ISL_425464\|20 | ···················· | ···················· | ···················· | ···················· | ···················· | ···················· |
| A | 2413 hCoV-19/England/NOTT-10DEDB/2020\|EPI_ISL_425465\|20 | ···················· | ···················· | ···················· | ···················· | ···················· | ···················· |
| A | 2414 hCoV-19/England/NOTT-10DEEA/2020\|EPI_ISL_425466\|20 | ···················· | ···················· | ···················· | ···················· | ···················· | ···················· |
| A | 2415 hCoV-19/England/NOTT-10DF05/2020\|EPI_ISL_425468\|20 | ···················· | ···················· | ···················· | ···················· | ···················· | ···················· |
| A | 2416 hCoV-19/England/NOTT-10DF23/2020\|EPI_ISL_425470\|20 | ···················· | ···················· | ···················· | ···················· | ···················· | ···················· |
| A | 2417 hCoV-19/England/NOTT-10DF32/2020\|EPI_ISL_425471\|20 | ···················· | ···················· | ···················· | ···················· | ···················· | ···················· |
| A | 2418 hCoV-19/England/NOTT-10DF41/2020\|EPI_ISL_425472\|20 | ···················· | ···················· | ···················· | ···················· | ···················· | ···················· |
| A | 2419 hCoV-19/England/NOTT-10DF50/2020\|EPI_ISL_425473\|20 | ···················· | ···················· | ···················· | ···················· | ···················· | ···················· |
| A | 2420 hCoV-19/England/NOTT-10DF6F/2020\|EPI_ISL_425474\|20 | ···················· | ···················· | ···················· | ···················· | ···················· | ···················· |
| A | 2421 hCoV-19/England/NOTT-10DF7E/2020\|EPI_ISL_425475\|20 | ···················· | ···················· | ···················· | ···················· | ···················· | ···················· |
| A | 2422 hCoV-19/England/NOTT-10DF9C/2020\|EPI_ISL_425476\|20 | ···················· | ···················· | ···················· | ···················· | ···················· | ···················· |
| A | 2423 hCoV-19/England/NOTT-10DFAB/2020\|EPI_ISL_425477\|20 | ···················· | ···················· | ···················· | ···················· | ···················· | ···················· |
| A | 2424 hCoV-19/England/NOTT-10DFC9/2020\|EPI_ISL_425479\|20 | ···················· | ···················· | ···················· | ···················· | ···················· | ···················· |
| A | 2425 hCoV-19/England/NOTT-10DFD8/2020\|EPI_ISL_425480\|20 | ···················· | ···················· | ···················· | ···················· | ···················· | ···················· |
| A | 2426 hCoV-19/England/NOTT-10DFE7/2020\|EPI_ISL_425481\|20 | ···················· | ···················· | ···················· | ···················· | ···················· | ···················· |
| A | 2427 hCoV-19/England/NOTT-10DFF6/2020\|EPI_ISL_425482\|20 | ···················· | ···················· | ···················· | ···················· | ···················· | ···················· |
| A | 2428 hCoV-19/England/NOTT-10E001/2020\|EPI_ISL_425483\|20 | ···················· | ···················· | ···················· | ···················· | ···················· | ···················· |
| A | 2429 hCoV-19/England/NOTT-10E010/2020\|EPI_ISL_425484\|20 | ···················· | ···················· | ···················· | ···················· | ···················· | ···················· |
| A | 2430 hCoV-19/England/NOTT-10E02F/2020\|EPI_ISL_425485\|20 | ···················· | ···················· | ···················· | ···················· | ···················· | ···················· |
| A | 2431 hCoV-19/England/NOTT-10E03E/2020\|EPI_ISL_425486\|20 | ···················· | ···················· | ···················· | ···················· | ···················· | ···················· |
| A | 2432 hCoV-19/England/NOTT-10E04D/2020\|EPI_ISL_425487\|20 | ···················· | ···················· | ···················· | ···················· | ···················· | ···················· |
| A | 2433 hCoV-19/England/NOTT-10E05C/2020\|EPI_ISL_425488\|20 | ···················· | ···················· | ···················· | ···················· | ···················· | ···················· |
| A | 2434 hCoV-19/England/NOTT-10E06B/2020\|EPI_ISL_425489\|20 | ···················· | ···················· | ···················· | ···················· | ···················· | ···················· |

| | | | | | | | |
|---|---|---|---|---|---|---|---|
| A | 2435 hCoV-19/England/NOTT-10E07A/2020\|EPI_ISL_425490\|20 | ···················· | ···················· | ···················· | ···················· | ···················· | ···················· |
| A | 2436 hCoV-19/England/NOTT-10E089/2020\|EPI_ISL_425491\|20 | ···················· | ···················· | ···················· | ···················· | ···················· | ···················· |
| A | 2437 hCoV-19/England/NOTT-10E098/2020\|EPI_ISL_425492\|20 | ···················· | ···················· | ···················· | ···················· | ···················· | ···················· |
| A | 2438 hCoV-19/England/NOTT-10E0A7/2020\|EPI_ISL_425493\|20 | ···················· | ···················· | ···················· | ···················· | ···················· | ···················· |
| A | 2439 hCoV-19/England/NOTT-10E0B6/2020\|EPI_ISL_425494\|20 | ···················· | ···················· | ···················· | ···················· | ···················· | ···················· |
| A | 2440 hCoV-19/England/NOTT-10E0C5/2020\|EPI_ISL_425495\|20 | ···················· | ···················· | ···················· | ···················· | ···················· | ···················· |
| A | 2441 hCoV-19/England/NOTT-10E0D4/2020\|EPI_ISL_425496\|20 | ···················· | ···················· | ···················· | ···················· | ···················· | ···················· |
| A | 2442 hCoV-19/England/NOTT-10E0E3/2020\|EPI_ISL_425497\|20 | ···················· | ···················· | ···················· | ···················· | ···················· | ···················· |
| A | 2443 hCoV-19/England/NOTT-10E0F2/2020\|EPI_ISL_425498\|20 | ···················· | ···················· | ···················· | ···················· | ···················· | ···················· |
| A | 2444 hCoV-19/Australia/VIC397/2020\|EPI_ISL_426700\|2020- | ···················· | ···················· | ···················· | ···················· | ···················· | ···················· |
| A | 2445 hCoV-19/Australia/VIC398/2020\|EPI_ISL_426701\|2020- | ···················· | ···················· | ···················· | ···················· | ···················· | ···················· |
| A | 2446 hCoV-19/Australia/VIC399/2020\|EPI_ISL_426702\|2020- | ···················· | ···················· | ···················· | ···················· | ···················· | ···················· |
| A | 2447 hCoV-19/Australia/VIC400/2020\|EPI_ISL_426703\|2020- | ···················· | ···················· | ···················· | ···················· | ···················· | ···················· |
| A | 2448 hCoV-19/Australia/VIC402/2020\|EPI_ISL_426705\|2020- | ···················· | ···················· | ···················· | ···················· | ···················· | ···················· |
| A | 2449 hCoV-19/Australia/VIC403/2020\|EPI_ISL_426706\|2020- | ···················· | ···················· | ···················· | ···················· | ···················· | ···················· |
| A | 2450 hCoV-19/Australia/VIC404/2020\|EPI_ISL_426707\|2020- | ···················· | ···················· | ···················· | ···················· | ···················· | ···················· |
| A | 2451 hCoV-19/Australia/VIC407/2020\|EPI_ISL_426709\|2020- | ···················· | ···················· | ···················· | ···················· | ···················· | ···················· |
| A | 2469 hCoV-19/Australia/VIC444/2020\|EPI_ISL_426741\|2020- | ···················· | ···················· | ···················· | ···················· | ···················· | ···················· |
| A | 2471 hCoV-19/Australia/VIC449/2020\|EPI_ISL_426745\|2020- | ···················· | ···················· | ···················· | ···················· | ···················· | ···················· |
| A | 2476 hCoV-19/Australia/VIC458/2020\|EPI_ISL_426752\|2020- | ···················· | ···················· | ···················· | ···················· | ···················· | ···················· |
| A | 2481 hCoV-19/Australia/VIC471/2020\|EPI_ISL_426763\|2020- | ···················· | ···················· | ···················· | ···················· | ···················· | ···············R···· |
| A | 2482 hCoV-19/Australia/VIC492/2020\|EPI_ISL_426765\|2020- | ···················· | ·············A······ | ···················· | ···················· | ···················· | ···················· |
| A | 2484 hCoV-19/Australia/VIC494/2020\|EPI_ISL_426767\|2020- | ···················· | ···················· | ···················· | ···················· | ···················· | ···················· |
| A | 2487 hCoV-19/Australia/VIC497/2020\|EPI_ISL_426770\|2020- | ···················· | ···················· | ···················· | ···················· | ···················· | ···················· |
| A | 2488 hCoV-19/Australia/VIC498/2020\|EPI_ISL_426771\|2020- | ···················· | ···················· | ···················· | ···················· | ···················· | ···················· |
| A | 2489 hCoV-19/Australia/VIC500/2020\|EPI_ISL_426772\|2020- | ···················· | ···················· | ···················· | ···················· | ···················· | ···················· |
| A | 2490 hCoV-19/Australia/VIC502/2020\|EPI_ISL_426773\|2020- | ···················· | ···················· | ···················· | ···················· | ···················· | ···················· |
| A | 2492 hCoV-19/Australia/VIC504/2020\|EPI_ISL_426775\|2020- | ···················· | ···················· | ···················· | ···················· | ···················· | ···················· |
| A | 2493 hCoV-19/Australia/VIC506/2020\|EPI_ISL_426777\|2020- | ···················· | ···················· | ···················· | ···················· | ···················· | ···················· |
| A | 2494 hCoV-19/Australia/VIC508/2020\|EPI_ISL_426778\|2020- | ···················· | ···················· | ···················· | ···················· | ···················· | ···················· |
| A | 2495 hCoV-19/Australia/VIC509/2020\|EPI_ISL_426779\|2020- | ···················· | ···················· | ···················· | ···················· | ···················· | ···················· |
| A | 2496 hCoV-19/Sweden/RV-FOI-6/2020\|EPI_ISL_428201\|2020-0 | ···················· | ···················· | ···················· | ···················· | ···················· | ···················· |
| A | 2497 hCoV-19/Poland/Pom1/2020\|EPI_ISL_428209\|2020-04-06 | ···················· | ···················· | ···················· | ···················· | ···················· | ···················· |
| A | 2498 hCoV-19/USA/WI-UW-164/2020\|EPI_ISL_428260\|2020-03- | ···················· | ···················· | ···················· | ···················· | ···················· | ···················· |
| A | 2499 hCoV-19/USA/WI-UW-165/2020\|EPI_ISL_428261\|2020-03- | ···················· | ···················· | ···················· | ···················· | ···················· | ···················· |
| A | 2500 hCoV-19/USA/WI-UW-166/2020\|EPI_ISL_428262\|2020-03- | ···················· | ···················· | ···················· | ···················· | ···················· | ···················· |
| A | 2501 hCoV-19/USA/WI-UW-167/2020\|EPI_ISL_428263\|2020-03- | ···················· | ···················· | ···················· | ···················· | ···················· | ···················· |
| A | 2502 hCoV-19/USA/WI-UW-168/2020\|EPI_ISL_428264\|2020-03- | ···················· | ···················· | ···················· | ···················· | ···················· | ···················· |
| A | 2503 hCoV-19/USA/WI-UW-169/2020\|EPI_ISL_428265\|2020-03- | ···················· | ···················· | ···················· | ···················· | ···················· | ···················· |
| A | 2504 hCoV-19/USA/WI-UW-170/2020\|EPI_ISL_428266\|2020-03- | ···················· | ···················· | ···················· | ···················· | ···················· | ···················· |
| A | 2505 hCoV-19/USA/WI-UW-171/2020\|EPI_ISL_428267\|2020-03- | ···················· | ···················· | ···················· | ···················· | ···················· | ···················· |
| A | 2506 hCoV-19/USA/WI-UW-172/2020\|EPI_ISL_428268\|2020-03- | ···················· | ···················· | ···················· | ···················· | ···················· | ···················· |
| A | 2507 hCoV-19/USA/WI-UW-173/2020\|EPI_ISL_428269\|2020-03- | ···················· | ···················· | ···················· | ···················· | ···················· | ···················· |
| A | 2508 hCoV-19/USA/WI-UW-174/2020\|EPI_ISL_428270\|2020-03- | ···················· | ···················· | ···················· | ···················· | ···················· | ···················· |
| A | 2509 hCoV-19/USA/WI-UW-175/2020\|EPI_ISL_428271\|2020-03- | ···················· | ···················· | ···················· | ···················· | ···················· | ···················· |
| A | 2510 hCoV-19/USA/WI-UW-176/2020\|EPI_ISL_428272\|2020-03- | ···················· | ···················· | ···················· | ···················· | ···················· | ···················· |
| A | 2511 hCoV-19/USA/WI-UW-177/2020\|EPI_ISL_428273\|2020-03- | ···················· | ···················· | ···················· | ···················· | ···················· | ···················· |
| A | 2512 hCoV-19/USA/WI-UW-178/2020\|EPI_ISL_428274\|2020-03- | ···················· | ···················· | ···················· | ···················· | ···················· | ···················· |
| A | 2513 hCoV-19/USA/WI-UW-179/2020\|EPI_ISL_428275\|2020-03- | ···················· | ···················· | ···················· | ···················· | ···················· | ···················· |
| A | 2514 hCoV-19/USA/WI-UW-180/2020\|EPI_ISL_428276\|2020-03- | ···················· | ···················· | ···················· | ···················· | ···················· | ···················· |
| A | 2515 hCoV-19/USA/WI-UW-181/2020\|EPI_ISL_428277\|2020-03- | ···················· | ···················· | ···················· | ···················· | ···················· | ···················· |
| A | 2516 hCoV-19/USA/WI-UW-182/2020\|EPI_ISL_428278\|2020-03- | ···················· | ···················· | ···················· | ···················· | ···················· | ···················· |
| A | 2517 hCoV-19/USA/WI-UW-183/2020\|EPI_ISL_428279\|2020-03- | ···················· | ···················· | ···················· | ···················· | ···················· | ···················· |
| A | 2518 hCoV-19/USA/WI-UW-184/2020\|EPI_ISL_428280\|2020-03- | ···················· | ···················· | ···················· | ···················· | ···················· | ···················· |
| A | 2519 hCoV-19/USA/WI-UW-185/2020\|EPI_ISL_428281\|2020-03- | ···················· | ···················· | ···················· | ···················· | ···················· | ···················· |
| A | 2520 hCoV-19/USA/WI-UW-186/2020\|EPI_ISL_428282\|2020-03- | ···················· | ···················· | ···················· | ···················· | ···················· | ···················· |
| A | 2521 hCoV-19/USA/WI-UW-187/2020\|EPI_ISL_428283\|2020-03- | ···················· | ···················· | ···················· | ···················· | ···················· | ···················· |
| A | 2522 hCoV-19/USA/WI-UW-188/2020\|EPI_ISL_428284\|2020-03- | ···················· | ···················· | ···················· | ···················· | ···················· | ···················· |
| A | 2523 hCoV-19/USA/WI-UW-189/2020\|EPI_ISL_428285\|2020-03- | ···················· | ···················· | ···················· | ···················· | ···················· | ···················· |
| A | 2524 hCoV-19/USA/WI-UW-190/2020\|EPI_ISL_428286\|2020-03- | ···················· | ·········A·········· | ···················· | ···················· | ···················· | ···················· |
| A | 2525 hCoV-19/USA/WI-UW-191/2020\|EPI_ISL_428287\|2020-03- | ···················· | ···················· | ···················· | ···················· | ···················· | ···················· |
| A | 2526 hCoV-19/USA/WI-UW-192/2020\|EPI_ISL_428288\|2020-03- | ···················· | ···················· | ···················· | ···················· | ···················· | ···················· |
| A | 2527 hCoV-19/USA/WI-UW-193/2020\|EPI_ISL_428289\|2020-03- | ···················· | ···················· | ···················· | ···················· | ···················· | ···················· |
| A | 2528 hCoV-19/USA/WI-UW-194/2020\|EPI_ISL_428290\|2020-03- | ···················· | ···················· | ···················· | ···················· | ···················· | ···················· |
| A | 2529 hCoV-19/USA/WI-UW-195/2020\|EPI_ISL_428291\|2020-03- | ···················· | ···················· | ···················· | ···················· | ···················· | ···················· |
| A | 2530 hCoV-19/USA/WI-UW-196/2020\|EPI_ISL_428292\|2020-03- | ···················· | ···················· | ···················· | ···················· | ···················· | ···················· |
| A | 2531 hCoV-19/USA/WI-UW-197/2020\|EPI_ISL_428293\|2020-03- | ···················· | ···················· | ···················· | ···················· | ···················· | ···················· |
| A | 2532 hCoV-19/USA/WI-UW-198/2020\|EPI_ISL_428294\|2020-03- | ···················· | ···················· | ···················· | ···················· | ···················· | ···················· |
| A | 2533 hCoV-19/USA/WI-UW-199/2020\|EPI_ISL_428295\|2020-03- | ···················· | ···················· | ···················· | ···················· | ···················· | ···················· |
| A | 2534 hCoV-19/USA/WI-UW-200/2020\|EPI_ISL_428296\|2020-03- | ···················· | ···················· | ···················· | ···················· | ···················· | ···················· |
| A | 2535 hCoV-19/USA/WI-UW-201/2020\|EPI_ISL_428297\|2020-03- | ···················· | ···················· | ···················· | ···················· | ···················· | ···················· |
| A | 2536 hCoV-19/USA/WI-UW-202/2020\|EPI_ISL_428298\|2020-03- | ···················· | ···················· | ···················· | ···················· | ···················· | ···················· |
| A | 2537 hCoV-19/USA/WI-UW-203/2020\|EPI_ISL_428299\|2020-03- | ···················· | ···················· | ···················· | ···················· | ···················· | ···················· |
| A | 2538 hCoV-19/Denmark/ALAB-SSI412A/2020\|EPI_ISL_429508\|2 | ···················· | ···················· | ···················· | ···················· | ···················· | ···················· |
| A | 2539 hCoV-19/Denmark/ALAB-SSI413/2020\|EPI_ISL_429509\|20 | ···················· | ···················· | ···················· | ···················· | ···················· | ···················· |
| A | 2540 hCoV-19/Denmark/ALAB-SSI437/2020\|EPI_ISL_429530\|20 | ···················· | ···················· | ···················· | ···················· | ···················· | ···················· |
| A | 2541 hCoV-19/Denmark/ALAB-SSI444/2020\|EPI_ISL_429533\|20 | ···················· | ···················· | ···················· | ···················· | ···················· | ···················· |
| A | 2542 hCoV-19/Denmark/ALAB-SSI446/2020\|EPI_ISL_429534\|20 | ···················· | ···················· | ···················· | ···················· | ···················· | ···················· |

| | | | | | | | | | |
|---|---|---|---|---|---|---|---|---|---|
| A | 2543 | hCoV-19/Denmark/ALAB-SSI448/2020\|EPI_ISL_429535\|20 | .................... | .................... | .................... | .................... | .................... | .................... | |
| A | 2544 | hCoV-19/Denmark/ALAB-SSI449/2020\|EPI_ISL_429536\|20 | .................... | .................... | .................... | .................... | .................... | .................... | |
| A | 2545 | hCoV-19/Denmark/ALAB-SSI450/2020\|EPI_ISL_429537\|20 | .................... | .................... | .................... | .................... | .................... | .................... | |
| A | 2546 | hCoV-19/Denmark/ALAB-SSI451/2020\|EPI_ISL_429538\|20 | .................... | .................... | .................... | .................... | .................... | .................... | |
| A | 2547 | hCoV-19/Denmark/ALAB-SSI454/2020\|EPI_ISL_429539\|20 | .................... | .................... | .................... | .................... | .................... | .................... | |
| A | 2548 | hCoV-19/Denmark/ALAB-SSI456/2020\|EPI_ISL_429540\|20 | .................... | .................... | .................... | .................... | .................... | .................... | |
| A | 2549 | hCoV-19/Denmark/ALAB-SSI461/2020\|EPI_ISL_429542\|20 | .................... | .................... | .................... | .................... | .................... | .................... | |
| A | 2550 | hCoV-19/Denmark/ALAB-SSI462/2020\|EPI_ISL_429543\|20 | .................... | .................... | .................... | .................... | .................... | .................... | |
| A | 2551 | hCoV-19/Denmark/ALAB-SSI463/2020\|EPI_ISL_429544\|20 | .................... | .................... | .................... | .................... | .................... | .................... | |
| A | 2552 | hCoV-19/Denmark/ALAB-SSI464/2020\|EPI_ISL_429545\|20 | .................... | .................... | .................... | .................... | .................... | .................... | |
| A | 2553 | hCoV-19/Denmark/ALAB-SSI466/2020\|EPI_ISL_429546\|20 | .................... | .................... | .................... | .................... | .................... | .................... | |
| A | 2554 | hCoV-19/Denmark/ALAB-SSI467/2020\|EPI_ISL_429547\|20 | .................... | .................... | .................... | .................... | .................... | .................... | |
| A | 2555 | hCoV-19/Denmark/ALAB-SSI468/2020\|EPI_ISL_429548\|20 | .................... | .................... | .................... | .................... | .................... | .................... | |
| A | 2556 | hCoV-19/Denmark/ALAB-SSI469/2020\|EPI_ISL_429549\|20 | .................... | .................... | .................... | .................... | .................... | .................... | |
| A | 2557 | hCoV-19/Denmark/ALAB-SSI595/2020\|EPI_ISL_429590\|20 | .................... | .................... | .................... | .................... | .................... | .................... | |
| A | 2558 | hCoV-19/USA/WA-UW-4546/2020\|EPI_ISL_429598\|2020-03 | .................... | .................... | .................... | .................... | .................... | .................... | |
| A | 2559 | hCoV-19/Germany/HH-UMC-01/2020\|EPI_ISL_450413\|2020 | .................... | .................... | .................... | .................... | .................... | .................... | |
| A | 2560 | hCoV-19/Beijing/IME-HZ01/2020\|EPI_ISL_450444\|2020- | .................... | .................... | .................... | .................... | .................... | .................... | |
| A | 2561 | hCoV-19/Wuhan/WH03/2020\|EPI_ISL_406800\|2020-01-01 | .................... | .................... | .................... | .................... | .................... | .................... | |
| A | 2562 | hCoV-19/Wuhan/WH04/2020\|EPI_ISL_406801\|2020-01-05 | .................... | .................... | .................... | .................... | .................... | .................... | |
| A | 2563 | hCoV-19/USA/MN56-MDH56/2020\|EPI_ISL_417500\|2020-03 | .................... | .................... | .................... | .................... | .................... | .................... | |
| A | 2564 | hCoV-19/USA/MN57-MDH57/2020\|EPI_ISL_417501\|2020-03 | .................... | .................... | .................... | .................... | .................... | .................... | |
| A | 2565 | hCoV-19/USA/MN58-MDH58/2020\|EPI_ISL_417502\|2020-03 | .................... | .................... | .................... | .................... | .................... | .................... | |
| A | 2566 | hCoV-19/USA/MN59-MDH59/2020\|EPI_ISL_417503\|2020-03 | .................... | .................... | .................... | .................... | .................... | .................... | |
| A | 2567 | hCoV-19/USA/WI-UW-15/2020\|EPI_ISL_417504\|2020-03-1 | .................... | .................... | .................... | .................... | .................... | .................... | |
| A | 2568 | hCoV-19/USA/WI-UW-11/2020\|EPI_ISL_417505\|2020-03-1 | .................... | .................... | .................... | .................... | .................... | .................... | |
| A | 2569 | hCoV-19/USA/WI-UW-12/2020\|EPI_ISL_417506\|2020-03-1 | .................... | .................... | .................... | .................... | .................... | .................... | |
| A | 2570 | hCoV-19/Taiwan/CGMH-CGU-07/2020\|EPI_ISL_417520\|202 | .................... | .................... | .................... | .................... | .................... | .................... | |
| A | 2571 | hCoV-19/Taiwan/CGMH-CGU-08/2020\|EPI_ISL_417521\|202 | .................... | .................... | .................... | .................... | .................... | .................... | |
| A | 2572 | hCoV-19/Taiwan/CGMH-CGU-09/2020\|EPI_ISL_417522\|202 | .................... | .................... | .................... | .................... | .................... | .................... | |
| A | 2573 | hCoV-19/Taiwan/CGMH-CGU-10/2020\|EPI_ISL_417523\|202 | .................... | .................... | .................... | .................... | .................... | .................... | |
| A | 2574 | hCoV-19/Taiwan/CGMH-CGU-11/2020\|EPI_ISL_417524\|202 | .................... | .................... | .................... | .................... | .................... | .................... | |
| A | 2575 | hCoV-19/Luxembourg/LNS2848109/2020\|EPI_ISL_417527\| | .................... | .................... | .................... | .................... | .................... | .................... | |
| A | 2576 | hCoV-19/Luxembourg/LNS3694003/2020\|EPI_ISL_417528\| | .................... | .................... | .................... | .................... | .................... | .................... | |
| A | 2577 | hCoV-19/Iceland/184/2020\|EPI_ISL_417550\|2020-03-16 | .................... | .................... | .................... | .................... | .................... | .................... | |
| A | 2578 | hCoV-19/Iceland/187/2020\|EPI_ISL_417551\|2020-03-16 | .................... | .................... | .................... | .................... | .................... | .................... | |
| A | 2579 | hCoV-19/Iceland/2/2020\|EPI_ISL_417552\|2020-03-16 | .................... | .................... | .................... | .................... | .................... | .................... | |
| A | 2580 | hCoV-19/Iceland/220/2020\|EPI_ISL_417553\|2020-03-16 | .................... | .................... | .................... | .................... | .................... | .................... | |
| A | 2581 | hCoV-19/Iceland/227/2020\|EPI_ISL_417555\|2020-03-17 | .................... | .................... | .................... | .................... | .................... | .................... | |
| A | 2582 | hCoV-19/Iceland/228/2020\|EPI_ISL_417556\|2020-03-17 | .................... | .................... | .................... | .................... | .................... | .................... | |
| A | 2583 | hCoV-19/Iceland/229/2020\|EPI_ISL_417557\|2020-03-17 | .................... | .................... | .................... | .................... | .................... | .................... | |
| A | 2584 | hCoV-19/Iceland/230/2020\|EPI_ISL_417558\|2020-03-17 | .................... | .................... | .................... | .................... | .................... | .................... | |
| A | 2585 | hCoV-19/Iceland/242/2020\|EPI_ISL_417570\|2020-03-17 | .................... | .................... | .................... | .................... | .................... | .................... | |
| A | 2586 | hCoV-19/Iceland/243/2020\|EPI_ISL_417571\|2020-03-17 | .................... | .................... | .................... | .................... | .................... | .................... | |
| A | 2587 | hCoV-19/Iceland/244/2020\|EPI_ISL_417572\|2020-03-17 | .................... | .................... | .................... | .................... | .................... | .................... | |
| A | 2588 | hCoV-19/Iceland/245/2020\|EPI_ISL_417573\|2020-03-16 | .................... | .................... | .................... | .................... | .................... | .................... | |
| A | 2589 | hCoV-19/Iceland/246/2020\|EPI_ISL_417574\|2020-03-17 | .................... | .................... | .................... | .................... | .................... | .................... | |
| A | 2590 | hCoV-19/Iceland/247/2020\|EPI_ISL_417575\|2020-03-17 | .................... | .................... | .................... | .................... | .................... | .................... | |
| A | 2591 | hCoV-19/Iceland/248/2020\|EPI_ISL_417576\|2020-03-17 | .................... | .................... | .................... | .................... | .................... | .................... | |
| A | 2592 | hCoV-19/Iceland/249/2020\|EPI_ISL_417577\|2020-03-17 | .................... | .................... | .................... | .................... | .................... | .................... | |
| A | 2593 | hCoV-19/Iceland/252/2020\|EPI_ISL_417580\|2020-03-17 | .................... | .................... | .................... | .................... | .................... | .................... | |
| A | 2594 | hCoV-19/Iceland/253/2020\|EPI_ISL_417581\|2020-03-17 | .................... | .................... | .................... | .................... | .................... | .................... | |
| A | 2595 | hCoV-19/Iceland/254/2020\|EPI_ISL_417582\|2020-03-17 | .................... | .................... | .................... | .................... | .................... | .................... | |
| A | 2596 | hCoV-19/Iceland/255/2020\|EPI_ISL_417583\|2020-03-17 | .................... | .................... | .................... | .................... | .................... | .................... | |
| A | 2597 | hCoV-19/Iceland/256/2020\|EPI_ISL_417584\|2020-03-17 | .................... | .................... | .................... | .................... | .................... | .................... | |
| A | 2598 | hCoV-19/Iceland/257/2020\|EPI_ISL_417585\|2020-03-17 | .................... | .................... | .................... | .................... | .................... | .................... | |
| A | 2599 | hCoV-19/Iceland/258/2020\|EPI_ISL_417586\|2020-03-17 | .................... | .................... | .................... | .................... | .................... | .................... | |
| A | 2600 | hCoV-19/Iceland/259/2020\|EPI_ISL_417587\|2020-03-17 | .................... | .................... | .................... | .................... | .................... | .................... | |
| A | 2601 | hCoV-19/Iceland/260/2020\|EPI_ISL_417588\|2020-03-17 | .................... | .................... | .................... | .................... | .................... | .................... | |
| A | 2602 | hCoV-19/Iceland/261/2020\|EPI_ISL_417589\|2020-03-17 | .................... | .................... | .................... | .................... | .................... | .................... | |
| A | 2603 | hCoV-19/Iceland/262/2020\|EPI_ISL_417590\|2020-03-18 | .................... | .................... | .................... | .................... | .................... | .................... | |
| A | 2604 | hCoV-19/Iceland/263/2020\|EPI_ISL_417591\|2020-03-17 | .................... | .................... | .................... | .................... | .................... | .................... | |
| A | 2605 | hCoV-19/Iceland/264/2020\|EPI_ISL_417592\|2020-03-17 | .................... | ...................A | .................... | .................... | .................... | .................... | 1 |
| A | 2606 | hCoV-19/Iceland/267/2020\|EPI_ISL_417595\|2020-03-17 | .................... | .................... | .................... | .................... | .................... | .................... | |
| A | 2607 | hCoV-19/Iceland/268/2020\|EPI_ISL_417596\|2020-03-17 | .................... | .................... | .................... | .................... | .................... | .................... | |
| A | 2608 | hCoV-19/Iceland/269/2020\|EPI_ISL_417597\|2020-03-17 | .................... | .................... | .................... | .................... | .................... | .................... | |
| A | 2609 | hCoV-19/Iceland/270/2020\|EPI_ISL_417598\|2020-03-17 | .................... | .................... | .................... | .................... | .................... | .................... | |
| A | 2610 | hCoV-19/Iceland/271/2020\|EPI_ISL_417599\|2020-03-17 | .................... | .................... | .................... | .................... | .................... | .................... | |
| A | 2611 | hCoV-19/Belgium/CS-031052/2020\|EPI_ISL_418800\|2020 | .................... | .................... | .................... | .................... | .................... | .................... | |
| A | 2612 | hCoV-19/Australia/QLDID923/2020\|EPI_ISL_418801\|202 | .................... | .................... | .................... | .................... | .................... | .................... | |
| A | 2613 | hCoV-19/Australia/QLDID924/2020\|EPI_ISL_418802\|202 | .................... | .................... | .................... | .................... | .................... | .................... | |
| A | 2614 | hCoV-19/Australia/QLDID925/2020\|EPI_ISL_418803\|202 | .................... | .................... | .................... | .................... | .................... | .................... | |
| A | 2615 | hCoV-19/Australia/QLDID926/2020\|EPI_ISL_418804\|202 | .................... | .................... | .................... | .................... | .................... | .................... | |
| A | 2616 | hCoV-19/Belgium/VPE-030650/2020\|EPI_ISL_418805\|202 | .................... | .................... | .................... | .................... | .................... | .................... | |
| A | 2617 | hCoV-19/Belgium/BG-030551/2020\|EPI_ISL_418806\|2020 | .................... | .................... | .................... | .................... | .................... | .................... | |
| A | 2618 | hCoV-19/Australia/QLDID927/2020\|EPI_ISL_418807\|202 | .................... | .................... | .................... | .................... | .................... | .................... | |
| A | 2619 | hCoV-19/Australia/QLDID928/2020\|EPI_ISL_418808\|202 | .................... | .................... | .................... | .................... | .................... | .................... | |
| A | 2620 | hCoV-19/Japan/UT-NCGM02/2020\|EPI_ISL_418809\|2020-0 | .................... | .................... | .................... | .................... | .................... | .................... | |

| | | | | | | | | |
|---|---|---|---|---|---|---|---|---|
| A | 2621 | hCoV-19/Canada/NB_6/2020\|EPI_ISL_418811\|2020-03-12 | ···················· | ···················· | ···················· | ···················· | ···················· | ···················· |
| A | 2622 | hCoV-19/Canada/MB_8/2020\|EPI_ISL_418812\|2020-03-12 | ···················· | ···················· | ···················· | ···················· | ···················· | ···················· |
| A | 2623 | hCoV-19/Canada/MB_10/2020\|EPI_ISL_418813\|2020-03-1 | ···················· | ···················· | ···················· | ···················· | ···················· | ···················· |
| A | 2624 | hCoV-19/Canada/NS_13/2020\|EPI_ISL_418814\|2020-03-1 | ···················· | ···················· | ···················· | ···················· | ···················· | ···················· |
| A | 2625 | hCoV-19/Hong Kong/HKPU36-0702/2020\|EPI_ISL_418815\| | ···················· | ···················· | ···················· | ···················· | ···················· | ···················· |
| A | 2626 | hCoV-19/Canada/BC_0443574/2020\|EPI_ISL_418816\|2020 | ···················· | ···················· | ···················· | ···················· | ···················· | ···················· |
| A | 2627 | hCoV-19/Canada/BC_0554880/2020\|EPI_ISL_418817\|2020 | ···················· | ···················· | ···················· | ···················· | ···················· | ···················· |
| A | 2628 | hCoV-19/Canada/BC_1318414/2020\|EPI_ISL_418818\|2020 | ···················· | ···················· | ···················· | ···················· | ···················· | ···················· |
| A | 2629 | hCoV-19/Canada/BC_3808524/2020\|EPI_ISL_418819\|2020 | ···················· | ···················· | ···················· | ···················· | ···················· | ···················· |
| A | 2630 | hCoV-19/Canada/BC_3842755/2020\|EPI_ISL_418820\|2020 | ···················· | ···················· | ···················· | ···················· | ···················· | ···················· |
| A | 2631 | hCoV-19/Canada/BC_3968175/2020\|EPI_ISL_418821\|2020 | ···················· | ···················· | ···················· | ···················· | ···················· | ···················· |
| A | 2632 | hCoV-19/Canada/BC_3972884/2020\|EPI_ISL_418822\|2020 | ···················· | ···················· | ···················· | ···················· | ···················· | ···················· |
| A | 2633 | hCoV-19/Canada/BC_3989992/2020\|EPI_ISL_418823\|2020 | ···················· | ···················· | ···················· | ···················· | ···················· | ···················· |
| A | 2634 | hCoV-19/Canada/BC_4078583/2020\|EPI_ISL_418824\|2020 | ···················· | ···················· | ···················· | ···················· | ···················· | ···················· |
| A | 2635 | hCoV-19/Canada/BC_4118226/2020\|EPI_ISL_418825\|2020 | ···················· | ···················· | ···················· | ···················· | ···················· | ···················· |
| A | 2636 | hCoV-19/Canada/BC_4122951/2020\|EPI_ISL_418826\|2020 | ···················· | ···················· | ···················· | ···················· | ···················· | ···················· |
| A | 2637 | hCoV-19/Canada/BC_4540462/2020\|EPI_ISL_418829\|2020 | ···················· | ···················· | ···················· | ···················· | ···················· | ···················· |
| A | 2638 | hCoV-19/Canada/BC_4799711/2020\|EPI_ISL_418830\|2020 | ···················· | ···················· | ···················· | ···················· | ···················· | ···················· |
| A | 2639 | hCoV-19/Canada/BC_5275718/2020\|EPI_ISL_418831\|2020 | ···················· | ···················· | ···················· | ···················· | ···················· | ···················· |
| A | 2640 | hCoV-19/Canada/BC_5282984/2020\|EPI_ISL_418832\|2020 | ···················· | ···················· | ···················· | ···················· | ···················· | ···················· |
| A | 2641 | hCoV-19/Canada/BC_5306970/2020\|EPI_ISL_418833\|2020 | ···················· | ···················· | ···················· | ···················· | ···················· | ···················· |
| A | 2642 | hCoV-19/Canada/BC_5522039/2020\|EPI_ISL_418834\|2020 | ···················· | ···················· | ···················· | ···················· | ···················· | ···················· |
| A | 2643 | hCoV-19/Canada/BC_5979789/2020\|EPI_ISL_418835\|2020 | ···················· | ···················· | ···················· | ···················· | ···················· | ···················· |
| A | 2644 | hCoV-19/Canada/BC_6004567/2020\|EPI_ISL_418836\|2020 | ···················· | ···················· | ···················· | ···················· | ···················· | ···················· |
| A | 2645 | hCoV-19/Canada/BC_6129127/2020\|EPI_ISL_418837\|2020 | ···················· | ···················· | ···················· | ···················· | ···················· | ···················· |
| A | 2646 | hCoV-19/Canada/BC_8606204/2020\|EPI_ISL_418850\|2020 | ···················· | ···················· | ···················· | ···················· | ···················· | ···················· |
| A | 2647 | hCoV-19/Canada/BC_8622445/2020\|EPI_ISL_418851\|2020 | ···················· | ···················· | ···················· | ···················· | ···················· | ···················· |
| A | 2648 | hCoV-19/Canada/BC_8718874/2020\|EPI_ISL_418852\|2020 | ···················· | ···················· | ···················· | ···················· | ···················· | ···················· |
| A | 2649 | hCoV-19/Canada/BC_8894200/2020\|EPI_ISL_418853\|2020 | ···················· | ···················· | ···················· | ···················· | ···················· | ···················· |
| A | 2650 | hCoV-19/Canada/BC_8896915/2020\|EPI_ISL_418854\|2020 | ···················· | ···················· | ···················· | ···················· | ···················· | ···················· |
| A | 2651 | hCoV-19/Canada/BC_8897642/2020\|EPI_ISL_418855\|2020 | ···················· | ···················· | ···················· | ···················· | ···················· | ···················· |
| A | 2652 | hCoV-19/Canada/BC_9446031/2020\|EPI_ISL_418858\|2020 | ···················· | ···················· | ···················· | ···················· | ···················· | ···················· |
| A | 2653 | hCoV-19/USA/WA-UW313/2020\|EPI_ISL_418880\|2020-03-1 | ···················· | ···················· | ···················· | ···················· | ···················· | ···················· |
| A | 2654 | hCoV-19/USA/WA-UW314/2020\|EPI_ISL_418881\|2020-03-1 | ···················· | ···················· | ···················· | ···················· | ···················· | ···················· |
| A | 2655 | hCoV-19/USA/WA-UW315/2020\|EPI_ISL_418882\|2020-03-1 | ···················· | ···················· | ···················· | ···················· | ···················· | ···················· |
| A | 2656 | hCoV-19/USA/WA-UW316/2020\|EPI_ISL_418883\|2020-03-1 | ···················· | ···················· | ···················· | ···················· | ···················· | ···················· |
| A | 2657 | hCoV-19/USA/WA-UW317/2020\|EPI_ISL_418884\|2020-03-1 | ···················· | ···················· | ···················· | ···················· | ···················· | ···················· |
| A | 2658 | hCoV-19/USA/WA-UW319/2020\|EPI_ISL_418886\|2020-03-1 | ···················· | ···················· | ···················· | ···················· | ···················· | ···················· |
| A | 2659 | hCoV-19/USA/WA-UW320/2020\|EPI_ISL_418887\|2020-03-1 | ···················· | ···················· | ···················· | ···················· | ···················· | ···················· |
| A | 2660 | hCoV-19/USA/WA-UW321/2020\|EPI_ISL_418888\|2020-03-1 | ···················· | ···················· | ···················· | ···················· | ···················· | ···················· |
| A | 2661 | hCoV-19/USA/WA-UW322/2020\|EPI_ISL_418889\|2020-03-1 | ···················· | ···················· | ···················· | ···················· | ···················· | ···················· |
| A | 2662 | hCoV-19/Wales/PHWC-25779/2020\|EPI_ISL_422039\|2020- | ···················· | ····A··············· | ···················· | ···················· | ···················· | ···················· |
| A | 2663 | hCoV-19/Wales/PHWC-26D1C/2020\|EPI_ISL_422040\|2020- | ···················· | ····A··············· | ···················· | ···················· | ···················· | ···················· |
| A | 2664 | hCoV-19/Wales/PHWC-25C3E/2020\|EPI_ISL_422041\|2020- | ···················· | ···················· | ···················· | ···················· | ···················· | ···················· |
| A | 2665 | hCoV-19/Wales/PHWC-264BD/2020\|EPI_ISL_422042\|2020- | ···················· | ···················· | ···················· | ···················· | ···················· | ···················· |
| A | 2666 | hCoV-19/Wales/PHWC-269DC/2020\|EPI_ISL_422043\|2020- | ···················· | ···················· | ···················· | ···················· | ···················· | ···················· |
| A | 2667 | hCoV-19/Wales/PHWC-26E19/2020\|EPI_ISL_422044\|2020- | ···················· | ···················· | ···················· | ···················· | ···················· | ···················· |
| A | 2668 | hCoV-19/Wales/PHWC-25AF8/2020\|EPI_ISL_422045\|2020- | ···················· | ···················· | ···················· | ···················· | ···················· | ···················· |
| A | 2669 | hCoV-19/Wales/PHWC-2666C/2020\|EPI_ISL_422046\|2020- | ···················· | ···················· | ···················· | ···················· | ···················· | ···················· |
| A | 2670 | hCoV-19/Wales/PHWC-26F34/2020\|EPI_ISL_422047\|2020- | ···················· | ···················· | ···················· | ···················· | ···················· | ···················· |
| A | 2671 | hCoV-19/Wales/PHWC-26918/2020\|EPI_ISL_422048\|2020- | ···················· | ···················· | ···················· | ···················· | ···················· | ···················· |
| A | 2672 | hCoV-19/Wales/PHWC-2575B/2020\|EPI_ISL_422049\|2020- | ···················· | ····A··············· | ···················· | ···················· | ···················· | ···················· |
| A | 2673 | hCoV-19/Wales/PHWC-26DEF/2020\|EPI_ISL_422050\|2020- | ···················· | ····A··············· | ···················· | ···················· | ···················· | ···················· |
| A | 2674 | hCoV-19/Wales/PHWC-26A51/2020\|EPI_ISL_422051\|2020- | ···················· | ····A··············· | ···················· | ···················· | ···················· | ···················· |
| A | 2675 | hCoV-19/Wales/PHWC-26884/2020\|EPI_ISL_422052\|2020- | ···················· | ···················· | ···················· | ···················· | ···················· | ···················· |
| A | 2676 | hCoV-19/Wales/PHWC-25ABC/2020\|EPI_ISL_422053\|2020- | ···················· | ···················· | ···················· | ···················· | ···················· | ···················· |
| A | 2677 | hCoV-19/Wales/PHWC-26F9E/2020\|EPI_ISL_422054\|2020- | ···················· | ···················· | ···················· | ···················· | ···················· | ···················· |
| A | 2678 | hCoV-19/Wales/PHWC-25E47/2020\|EPI_ISL_422055\|2020- | ···················· | ····A··············· | ···················· | ···················· | ···················· | ···················· |
| A | 2679 | hCoV-19/Wales/PHWC-26CC4/2020\|EPI_ISL_422056\|2020- | ···················· | ····A··············· | ···················· | ···················· | ···················· | ···················· |
| A | 2680 | hCoV-19/Wales/PHWC-268A2/2020\|EPI_ISL_422057\|2020- | ···················· | ···················· | ···················· | ···················· | ···················· | ···················· |
| A | 2681 | hCoV-19/Wales/PHWC-26E91/2020\|EPI_ISL_422058\|2020- | ···················· | ···················· | ···················· | ···················· | ···················· | ···················· |
| A | 2682 | hCoV-19/Wales/PHWC-26FE9/2020\|EPI_ISL_422059\|2020- | ···················· | ···················· | ···················· | ···················· | ···················· | ···················· |
| A | 2683 | hCoV-19/Wales/PHWC-25FEA/2020\|EPI_ISL_422060\|2020- | ···················· | ···················· | ···················· | ···················· | ···················· | ···················· |
| A | 2684 | hCoV-19/Wales/PHWC-26E82/2020\|EPI_ISL_422061\|2020- | ···················· | ···················· | ···················· | ···················· | ···················· | ···················· |
| A | 2685 | hCoV-19/Wales/PHWC-269AF/2020\|EPI_ISL_422062\|2020- | ···················· | ·A·················· | ···················· | ···················· | ···················· | ···················· |
| A | 2686 | hCoV-19/Wales/PHWC-25B6E/2020\|EPI_ISL_422063\|2020- | ···················· | ···················· | ···················· | ···················· | ···················· | ···················· |
| A | 2687 | hCoV-19/Wales/PHWC-25C01/2020\|EPI_ISL_422064\|2020- | ···················· | ···················· | ···················· | ···················· | ···················· | ···················· |
| A | 2688 | hCoV-19/Wales/PHWC-2566D/2020\|EPI_ISL_422065\|2020- | ···················· | ···················· | ···················· | ···················· | ···················· | ···················· |
| A | 2689 | hCoV-19/Wales/PHWC-2573D/2020\|EPI_ISL_422066\|2020- | ···················· | ····A··············· | ···················· | ···················· | ···················· | ···················· |
| A | 2690 | hCoV-19/Wales/PHWC-25973/2020\|EPI_ISL_422067\|2020- | ···················· | ···················· | ···················· | ···················· | ···················· | ···················· |
| A | 2691 | hCoV-19/Wales/PHWC-26EFB/2020\|EPI_ISL_422068\|2020- | ···················· | ····A··············· | ···················· | ···················· | ···················· | ···················· |
| A | 2692 | hCoV-19/Wales/PHWC-270A9/2020\|EPI_ISL_422069\|2020- | ···················· | ···················· | ···················· | ···················· | ···················· | ···················· |
| A | 2693 | hCoV-19/Wales/PHWC-26839/2020\|EPI_ISL_422070\|2020- | ···················· | ···················· | ···················· | ···················· | ···················· | ···················· |
| A | 2694 | hCoV-19/Wales/PHWC-257B5/2020\|EPI_ISL_422071\|2020- | ···················· | ···················· | ···················· | ···················· | ···················· | ···················· |
| A | 2695 | hCoV-19/Wales/PHWC-26F52/2020\|EPI_ISL_422072\|2020- | ···················· | ···················· | ···················· | ···················· | ···················· | ···················· |
| A | 2696 | hCoV-19/Wales/PHWC-260AA/2020\|EPI_ISL_422073\|2020- | ···················· | ···················· | ···················· | ···················· | ···················· | ···················· |
| A | 2697 | hCoV-19/Wales/PHWC-26954/2020\|EPI_ISL_422074\|2020- | ···················· | ·A·················· | ···················· | ···················· | ···················· | ···················· |
| A | 2698 | hCoV-19/Wales/PHWC-26E46/2020\|EPI_ISL_422075\|2020- | ···················· | ···················· | ···················· | ···················· | ···················· | ···················· |

A 2699 hCoV-19/Wales/PHWC-26D3A/2020|EPI_ISL_422076|2020- ···················· ···················· ···················· ···················· ···················· ····················
A 2700 hCoV-19/Wales/PHWC-26A42/2020|EPI_ISL_422077|2020- ···················· ····A··············· ···················· ···················· ···················· ····················
A 2701 hCoV-19/Wales/PHWC-26013/2020|EPI_ISL_422078|2020- ···················· ····A··············· ···················· ···················· ···················· ····················
A 2702 hCoV-19/Wales/PHWC-26E37/2020|EPI_ISL_422079|2020- ···················· ···················· ···················· ···················· ···················· ····················
A 2703 hCoV-19/Wales/PHWC-26E55/2020|EPI_ISL_422080|2020- ···················· ···················· ···················· ···················· ···················· ····················
A 2704 hCoV-19/Wales/PHWC-26EBF/2020|EPI_ISL_422081|2020- ···················· ···················· ···················· ···················· ···················· ····················
A 2705 hCoV-19/Wales/PHWC-26BD6/2020|EPI_ISL_422082|2020- ···················· ···················· ···················· ···················· ···················· ····················
A 2706 hCoV-19/Wales/PHWC-2624A/2020|EPI_ISL_422083|2020- ···················· ···················· ···················· ···················· ···················· ····················
A 2707 hCoV-19/Wales/PHWC-26DB2/2020|EPI_ISL_422084|2020- ···················· ···················· ···················· ···················· ···················· ····················
A 2708 hCoV-19/Wales/PHWC-25FAE/2020|EPI_ISL_422085|2020- ···················· ···················· ···················· ···················· ···················· ····················
A 2709 hCoV-19/Wales/PHWC-2708B/2020|EPI_ISL_422086|2020- ···················· ···················· ···················· ···················· ···················· ····················
A 2710 hCoV-19/Wales/PHWC-26CA6/2020|EPI_ISL_422087|2020- ···················· ····A··············· ···················· ···················· ···················· ····················
A 2711 hCoV-19/Wales/PHWC-26DC1/2020|EPI_ISL_422088|2020- ···················· ···················· ···················· ···················· ···················· ····················
A 2712 hCoV-19/Wales/PHWC-26DA3/2020|EPI_ISL_422089|2020- ···················· ····A··············· ···················· ···················· ···················· ····················
A 2713 hCoV-19/Wales/PHWC-26ECE/2020|EPI_ISL_422090|2020- ···················· ···················· ···················· ···················· ···················· ····················
A 2714 hCoV-19/Wales/PHWC-264DB/2020|EPI_ISL_422091|2020- ···················· ···················· ···················· ···················· ···················· ····················
A 2715 hCoV-19/Wales/PHWC-26857/2020|EPI_ISL_422092|2020- ···················· ···················· ···················· ···················· ···················· ····················
A 2716 hCoV-19/Wales/PHWC-2707C/2020|EPI_ISL_422093|2020- ···················· ···················· ···················· ···················· ···················· ····················
A 2717 hCoV-19/Wales/PHWC-2617A/2020|EPI_ISL_422094|2020- ···················· ···················· ···················· ···················· ···················· ····················
A 2718 hCoV-19/Wales/PHWC-2705E/2020|EPI_ISL_422095|2020- ···················· ···················· ···················· ···················· ···················· ····················
A 2719 hCoV-19/Wales/PHWC-2675A/2020|EPI_ISL_422096|2020- ···················· ···················· ···················· ···················· ···················· ····················
A 2720 hCoV-19/Wales/PHWC-26D0D/2020|EPI_ISL_422097|2020- ···················· ···················· ···················· ···················· ···················· ····················
A 2721 hCoV-19/Wales/PHWC-26848/2020|EPI_ISL_422098|2020- ···················· ···················· ···················· ···················· ···················· ····················
A 2722 hCoV-19/Wales/PHWC-27021/2020|EPI_ISL_422099|2020- ···················· ···················· ···················· ···················· ···················· ····················
A 2723 hCoV-19/England/20140001704/2020|EPI_ISL_423370|20 ···················· ···················· ···················· ···················· ···················· ····················
A 2724 hCoV-19/England/20140002204/2020|EPI_ISL_423371|20 ···················· ···················· ···················· ···················· ···················· ····················
A 2725 hCoV-19/England/20140002804/2020|EPI_ISL_423373|20 ···················· ···················· ···················· ···················· ···················· ····················
A 2726 hCoV-19/England/20140003402/2020|EPI_ISL_423375|20 ···················· ···················· ···················· ···················· ···················· ····················
A 2727 hCoV-19/England/20140003404/2020|EPI_ISL_423376|20 ···················· ···················· ···················· ···················· ···················· ····N···············
A 2728 hCoV-19/England/20140004004/2020|EPI_ISL_423379|20 ···················· ···················· ···················· ···················· ···················· ····················
A 2730 hCoV-19/England/20140006204/2020|EPI_ISL_423381|20 ···················· ···················· ···················· ···················· ···················· ····················
A 2731 hCoV-19/England/20140007004/2020|EPI_ISL_423382|20 ···················· ···················· ···················· ···················· ···················· ····················
A 2732 hCoV-19/England/20140007602/2020|EPI_ISL_423383|20 ···················· ···················· ···················· ···················· ···················· ····················
A 2734 hCoV-19/England/20140009204/2020|EPI_ISL_423385|20 ···················· ···················· ···················· ···················· ···················· ····················
A 2736 hCoV-19/England/20140009404/2020|EPI_ISL_423387|20 ···················· ···················· ···················· ···················· ···················· ····················
A 2738 hCoV-19/England/20140064004/2020|EPI_ISL_423393|20 ···················· ···················· ···················· ···················· ···················· ····················
A 2739 hCoV-19/England/20140064504/2020|EPI_ISL_423394|20 ···················· ···················· ···················· ···················· ···················· ····················
A 2740 hCoV-19/England/20140064704/2020|EPI_ISL_423395|20 ···················· ···················· ···················· ···················· ···················· ····················
A 2741 hCoV-19/England/20140064804/2020|EPI_ISL_423396|20 ···················· ···················· ···················· ···················· ···················· ····················
A 2742 hCoV-19/England/20140066004/2020|EPI_ISL_423398|20 ···················· ···················· ···················· ···················· ···················· ····················
A 2743 hCoV-19/England/20140066204/2020|EPI_ISL_423399|20 ···················· ···················· ···················· ···················· ···················· ····················
A 2744 hCoV-19/Taiwan/TSGH-12/2020|EPI_ISL_428229|2020-03 ···················· ···················· ···················· ···················· ···················· ····················
A 2745 hCoV-19/Taiwan/TSGH-13/2020|EPI_ISL_428230|2020-03 ···················· ···················· ···················· ···················· ···················· ····················
A 2746 hCoV-19/Taiwan/TSGH-14/2020|EPI_ISL_428231|2020-03 ···················· ···················· ···················· ···················· ···················· ····················
A 2747 hCoV-19/USA/WI-UW-156/2020|EPI_ISL_428252|2020-03- ···················· ···················· ···················· ···················· ···················· ····················
A 2748 hCoV-19/USA/WI-UW-157/2020|EPI_ISL_428253|2020-03- ···················· ···················· ···················· ···················· ···················· ····················
A 2749 hCoV-19/USA/WI-UW-158/2020|EPI_ISL_428254|2020-03- ···················· ···················· ···················· ···················· ···················· ····················
A 2750 hCoV-19/USA/WI-UW-159/2020|EPI_ISL_428255|2020-03- ···················· ···················· ···················· ···················· ···················· ····················
A 2751 hCoV-19/USA/WI-UW-160/2020|EPI_ISL_428256|2020-03- ···················· ···················· ···················· ···················· ···················· ····················
A 2752 hCoV-19/USA/WI-UW-161/2020|EPI_ISL_428257|2020-03- ···················· ···················· ···················· ···················· ···················· ····················
A 2753 hCoV-19/USA/WI-UW-162/2020|EPI_ISL_428258|2020-03- ···················· ···················· ···················· ···················· ···················· ····················
A 2754 hCoV-19/USA/WI-UW-163/2020|EPI_ISL_428259|2020-03- ···················· ···················· ···················· ···················· ···················· ····················
A 2755 hCoV-19/Denmark/ALAB-SSI407/2020|EPI_ISL_429501|20 ···················· ···················· ···················· ···················· ···················· ····················
A 2756 hCoV-19/Denmark/ALAB-SSI408/2020|EPI_ISL_429502|20 ···················· ···················· ···················· ···················· ···················· ····················
A 2757 hCoV-19/Denmark/ALAB-SSI409/2020|EPI_ISL_429503|20 ···················· ···················· ···················· ···················· ···················· ····················
A 2758 hCoV-19/Denmark/ALAB-SSI411/2020|EPI_ISL_429505|20 ···················· ···················· ···················· ···················· ···················· ····················
A 2759 hCoV-19/Denmark/ALAB-SSI411A/2020|EPI_ISL_429506|2 ···················· ···················· ···················· ···················· ···················· ····················
A 2760 hCoV-19/Denmark/ALAB-SSI412/2020|EPI_ISL_429507|20 ···················· ···················· ···················· ···················· ···················· ····················
A 2761 hCoV-19/Denmark/ALAB-SSI413A/2020|EPI_ISL_429510|2 ···················· ···················· ···················· ···················· ···················· ····················
A 2762 hCoV-19/Denmark/ALAB-SSI414/2020|EPI_ISL_429511|20 ···················· ···················· ···················· ···················· ···················· ····················
A 2763 hCoV-19/Denmark/ALAB-SSI414A/2020|EPI_ISL_429512|2 ···················· ···················· ···················· ···················· ···················· ····················
A 2764 hCoV-19/Denmark/ALAB-SSI416/2020|EPI_ISL_429513|20 ···················· ···················· ···················· ···················· ···················· ····················
A 2765 hCoV-19/Denmark/ALAB-SSI417/2020|EPI_ISL_429514|20 ···················· ···················· ···················· ···················· ···················· ····················
A 2766 hCoV-19/Denmark/ALAB-SSI418/2020|EPI_ISL_429516|20 ···················· ···················· ···················· ···················· ···················· ····················
A 2767 hCoV-19/Denmark/ALAB-SSI419/2020|EPI_ISL_429517|20 ···················· ···················· ···················· ···················· ···················· ····················
A 2768 hCoV-19/Denmark/ALAB-SSI420/2020|EPI_ISL_429518|20 ···················· ···················· ···················· ···················· ···················· ····················
A 2769 hCoV-19/Denmark/ALAB-SSI420A/2020|EPI_ISL_429519|2 ···················· ···················· ···················· ···················· ···················· ····················
A 2770 hCoV-19/Denmark/ALAB-SSI423/2020|EPI_ISL_429520|20 ···················· ···················· ···················· ···················· ···················· ····················
A 2771 hCoV-19/Denmark/ALAB-SSI424/2020|EPI_ISL_429521|20 ···················· ···················· ···················· ···················· ···················· ····················
A 2772 hCoV-19/Denmark/ALAB-SSI425A/2020|EPI_ISL_429524|2 ···················· ···················· ···················· ···················· ···················· ····················
A 2773 hCoV-19/Denmark/ALAB-SSI426/2020|EPI_ISL_429525|20 ···················· ···················· ···················· ···················· ···················· ····················
A 2774 hCoV-19/Denmark/ALAB-SSI430/2020|EPI_ISL_429526|20 ···················· ···················· ···················· ···················· ···················· ····················
A 2775 hCoV-19/Denmark/ALAB-SSI431/2020|EPI_ISL_429527|20 ···················· ···················· ···················· ···················· ···················· ····················
A 2776 hCoV-19/Denmark/ALAB-SSI432/2020|EPI_ISL_429528|20 ···················· ···················· ···················· ···················· ···················· ····················
A 2777 hCoV-19/Denmark/ALAB-SSI434/2020|EPI_ISL_429529|20 ···················· ···················· ···················· ···················· ···················· ····················
A 2778 hCoV-19/Denmark/ALAB-SSI470/2020|EPI_ISL_429550|20 ···················· ···················· ···················· ···················· ···················· ····················
A 2779 hCoV-19/Denmark/ALAB-SSI471/2020|EPI_ISL_429551|20 ···················· ···················· ···················· ···················· ···················· ····················
A 2780 hCoV-19/Denmark/ALAB-SSI472/2020|EPI_ISL_429552|20 ···················· ···················· ···················· ···················· ···················· ····················

| | | | | | | | |
|---|---|---|---|---|---|---|---|
| A | 2781 hCoV-19/Denmark/ALAB-SSI473/2020\|EPI_ISL_429553\|20 | .................... | .................... | .................... | .................... | .................... | .................... |
| A | 2782 hCoV-19/Denmark/ALAB-SSI474/2020\|EPI_ISL_429554\|20 | .................... | .................... | .................... | .................... | .................... | .................... |
| A | 2783 hCoV-19/Denmark/ALAB-SSI475/2020\|EPI_ISL_429555\|20 | .................... | .................... | .................... | .................... | .................... | .................... |
| A | 2784 hCoV-19/Denmark/ALAB-SSI477/2020\|EPI_ISL_429556\|20 | .................... | .................... | .................... | .................... | .................... | .................... |
| A | 2785 hCoV-19/Denmark/ALAB-SSI478/2020\|EPI_ISL_429557\|20 | .................... | .................... | .................... | .................... | .................... | .................... |
| A | 2786 hCoV-19/Denmark/ALAB-SSI479/2020\|EPI_ISL_429558\|20 | .................... | .................... | .................... | .................... | .................... | .................... |
| A | 2787 hCoV-19/Denmark/ALAB-SSI480/2020\|EPI_ISL_429559\|20 | .................... | .................... | .................... | .................... | .................... | .................... |
| A | 2788 hCoV-19/Denmark/ALAB-SSI481/2020\|EPI_ISL_429560\|20 | .................... | .................... | .................... | .................... | .................... | .................... |
| A | 2789 hCoV-19/Denmark/ALAB-SSI482/2020\|EPI_ISL_429561\|20 | .................... | .................... | .................... | .................... | .................... | .................... |
| A | 2790 hCoV-19/Denmark/ALAB-SSI483/2020\|EPI_ISL_429562\|20 | .................... | .................... | .................... | .................... | .................... | .................... |
| A | 2791 hCoV-19/Denmark/ALAB-SSI484/2020\|EPI_ISL_429563\|20 | .................... | .................... | .................... | .................... | .................... | .................... |
| A | 2792 hCoV-19/Denmark/ALAB-SSI486/2020\|EPI_ISL_429564\|20 | .................... | .................... | .................... | .................... | .................... | .................... |
| A | 2793 hCoV-19/Denmark/ALAB-SSI488/2020\|EPI_ISL_429565\|20 | .................... | .................... | .................... | .................... | .................... | .................... |
| A | 2794 hCoV-19/Denmark/ALAB-SSI489/2020\|EPI_ISL_429566\|20 | .................... | .................... | .................... | .................... | .................... | .................... |
| A | 2795 hCoV-19/Denmark/ALAB-SSI490/2020\|EPI_ISL_429567\|20 | .................... | .................... | .................... | .................... | .................... | .................... |
| A | 2796 hCoV-19/Denmark/ALAB-SSI494/2020\|EPI_ISL_429569\|20 | .................... | .................... | .................... | .................... | .................... | .................... |
| A | 2797 hCoV-19/Denmark/ALAB-SSI495/2020\|EPI_ISL_429570\|20 | .................... | .................... | .................... | .................... | .................... | .................... |
| A | 2798 hCoV-19/Denmark/ALAB-SSI497/2020\|EPI_ISL_429571\|20 | .................... | .................... | .................... | .................... | .................... | .................... |
| A | 2799 hCoV-19/Denmark/ALAB-SSI501/2020\|EPI_ISL_429572\|20 | .................... | .................... | .................... | .................... | .................... | .................... |
| A | 2800 hCoV-19/Denmark/ALAB-SSI502/2020\|EPI_ISL_429573\|20 | .................... | .................... | .................... | .................... | .................... | .................... |
| A | 2801 hCoV-19/Denmark/ALAB-SSI506/2020\|EPI_ISL_429574\|20 | .................... | .................... | .................... | .................... | .................... | .................... |
| A | 2802 hCoV-19/Denmark/ALAB-SSI507/2020\|EPI_ISL_429575\|20 | .................... | .................... | .................... | .................... | .................... | .................... |
| A | 2803 hCoV-19/Denmark/ALAB-SSI509/2020\|EPI_ISL_429577\|20 | .................... | .................... | .................... | .................... | .................... | .................... |
| A | 2804 hCoV-19/Denmark/ALAB-SSI510/2020\|EPI_ISL_429578\|20 | .................... | .................... | .................... | .................... | .................... | .................... |
| A | 2805 hCoV-19/Denmark/ALAB-SSI511/2020\|EPI_ISL_429579\|20 | .................... | .................... | .................... | .................... | .................... | .................... |
| A | 2806 hCoV-19/Denmark/ALAB-SSI512/2020\|EPI_ISL_429580\|20 | .................... | .................... | .................... | .................... | .................... | .................... |
| A | 2807 hCoV-19/Denmark/ALAB-SSI513/2020\|EPI_ISL_429581\|20 | .................... | .................... | .................... | .................... | .................... | .................... |
| A | 2808 hCoV-19/Denmark/ALAB-SSI514/2020\|EPI_ISL_429582\|20 | .................... | .................... | .................... | .................... | .................... | .................... |
| A | 2809 hCoV-19/Denmark/ALAB-SSI515/2020\|EPI_ISL_429583\|20 | .................... | .................... | .................... | .................... | .................... | .................... |
| A | 2810 hCoV-19/Denmark/ALAB-SSI518/2020\|EPI_ISL_429584\|20 | .................... | .................... | .................... | .................... | .................... | .................... |
| A | 2811 hCoV-19/Denmark/ALAB-SSI520/2020\|EPI_ISL_429585\|20 | .................... | .................... | .................... | .................... | .................... | .................... |
| A | 2812 hCoV-19/Denmark/ALAB-SSI521/2020\|EPI_ISL_429586\|20 | .................... | .................... | .................... | .................... | .................... | .................... |
| A | 2813 hCoV-19/Denmark/ALAB-SSI522/2020\|EPI_ISL_429587\|20 | .................... | .................... | .................... | .................... | .................... | .................... |
| A | 2814 hCoV-19/Denmark/ALAB-SSI523/2020\|EPI_ISL_429588\|20 | .................... | .................... | .................... | .................... | .................... | .................... |
| A | 2815 hCoV-19/Denmark/ALAB-SSI592/2020\|EPI_ISL_429589\|20 | .................... | .................... | .................... | .................... | .................... | .................... |
| A | 2816 hCoV-19/Beijing/Beijing-01/2020\|EPI_ISL_514752\|202 | .................... | .................... | .................... | .................... | .................... | .................... |
| A | 2817 hCoV-19/Germany/BY-ChVir-929/2020\|EPI_ISL_406862\|2 | .................... | .................... | .................... | .................... | .................... | .................... |
| A | 2818 hCoV-19/USA/MA1/2020\|EPI_ISL_409067\|2020-01-29 | .................... | .A.................. | .................... | .................... | .................... | .................... |
| A | 2819 hCoV-19/USA/CA-CDC-6/2020\|EPI_ISL_410044\|2020-01-2 | .................... | .................... | .................... | .................... | .................... | .................... |
| A | 2820 hCoV-19/USA/IL2/2020\|EPI_ISL_410045\|2020-01-28 | .................... | .................... | .................... | .................... | .................... | .................... |
| A | 2821 hCoV-19/USA/CA-CDC-9/2020\|EPI_ISL_412862\|2020-02-2 | .................... | .................... | .................... | .................... | .................... | .................... |
| A | 2822 hCoV-19/South Korea/KCDC05/2020\|EPI_ISL_412869\|202 | .................... | .................... | .................... | .................... | .................... | .................... |
| A | 2823 hCoV-19/South Korea/KCDC06/2020\|EPI_ISL_412870\|202 | .................... | .................... | .................... | .................... | .................... | .................... |
| A | 2824 hCoV-19/South Korea/KCDC07/2020\|EPI_ISL_412871\|202 | .................... | .................... | .................... | .................... | .................... | .................... |
| A | 2825 hCoV-19/South Korea/KCDC12/2020\|EPI_ISL_412872\|202 | .................... | .................... | .................... | .................... | .................... | .................... |
| A | 2826 hCoV-19/South Korea/KCDC24/2020\|EPI_ISL_412873\|202 | .................... | .................... | .................... | .................... | .................... | .................... |
| A | 2827 hCoV-19/Switzerland/TI-SNRCI-29919486/2020\|EPI_ISL | .................... | .................... | .................... | .................... | .................... | .................... |
| A | 2828 hCoV-19/Switzerland/GE-SNRCI-29933895/2020\|EPI_ISL | .................... | .................... | .................... | .................... | .................... | .................... |
| A | 2829 hCoV-19/Switzerland/AG-SNRCI-29940361/2020\|EPI_ISL | .................... | .................... | .................... | .................... | .................... | .................... |
| A | 2830 hCoV-19/Shanghai/SH0022/2020\|EPI_ISL_416331\|2020-0 | .................... | .................... | .................... | .................... | .................... | .................... |
| A | 2831 hCoV-19/USA/WI-UW-23/2020\|EPI_ISL_417507\|2020-03-1 | .................... | .................... | .................... | .................... | .................... | .................... |
| A | 2832 hCoV-19/USA/WI-UW-21/2020\|EPI_ISL_417508\|2020-03-1 | .................... | .................... | .................... | .................... | .................... | .................... |
| A | 2833 hCoV-19/USA/WI-UW-16/2020\|EPI_ISL_417509\|2020-03-1 | .................... | .................... | .................... | .................... | .................... | .................... |
| A | 2834 hCoV-19/USA/WI-UW-19/2020\|EPI_ISL_417510\|2020-03-1 | .................... | .................... | .................... | .................... | .................... | .................... |
| A | 2835 hCoV-19/USA/WI-UW-24/2020\|EPI_ISL_417512\|2020-03-1 | .................... | .................... | .................... | .................... | .................... | .................... |
| A | 2836 hCoV-19/USA/WI-UW-14/2020\|EPI_ISL_417513\|2020-03-1 | .................... | .................... | .................... | .................... | .................... | .................... |
| A | 2837 hCoV-19/USA/WI-UW-22/2020\|EPI_ISL_417514\|2020-03-1 | .................... | .................... | .................... | .................... | .................... | .................... |
| A | 2838 hCoV-19/USA/WI-UW-18/2020\|EPI_ISL_417515\|2020-03-1 | .................... | .................... | .................... | .................... | .................... | .................... |
| A | 2839 hCoV-19/USA/WI-UW-13/2020\|EPI_ISL_417516\|2020-03-1 | .................... | .................... | .................... | .................... | .................... | .................... |
| A | 2840 hCoV-19/USA/WI-UW-17/2020\|EPI_ISL_417517\|2020-03-1 | .................... | .................... | .................... | .................... | .................... | .................... |
| A | 2841 hCoV-19/Taiwan/CGMH-CGU-06/2020\|EPI_ISL_417519\|202 | .................... | .................... | .................... | .................... | .................... | .................... |
| A | 2842 hCoV-19/Taiwan/CGMH-CGU-12/2020\|EPI_ISL_417525\|202 | .................... | .................... | .................... | .................... | .................... | .................... |
| A | 2843 hCoV-19/Luxembourg/LNS9371718/2020\|EPI_ISL_417526\| | .................... | .................... | .................... | .................... | .................... | .................... |
| A | 2845 hCoV-19/Iceland/334/2020\|EPI_ISL_417663\|2020-03-14 | .................... | .................... | .................... | .................... | .................... | .................... |
| A | 2846 hCoV-19/Canada/BC_6404889/2020\|EPI_ISL_418838\|2020 | .................... | .................... | .................... | .................... | .................... | .................... |
| A | 2847 hCoV-19/Canada/BC_6489864/2020\|EPI_ISL_418839\|2020 | .................... | .................... | .................... | .................... | .................... | .................... |
| A | 2848 hCoV-19/Canada/BC_6502001/2020\|EPI_ISL_418840\|2020 | .................... | .................... | .................... | .................... | .................... | .................... |
| A | 2849 hCoV-19/Canada/BC_6981299/2020\|EPI_ISL_418841\|2020 | .................... | .................... | .................... | .................... | .................... | .................... |
| A | 2850 hCoV-19/Canada/BC_6997898/2020\|EPI_ISL_418842\|2020 | .................... | .................... | .................... | .................... | .................... | .................... |
| A | 2851 hCoV-19/Canada/BC_7277381/2020\|EPI_ISL_418843\|2020 | .................... | .................... | .................... | .................... | .................... | .................... |
| A | 2852 hCoV-19/Canada/BC_7493713/2020\|EPI_ISL_418844\|2020 | .................... | .................... | .................... | .................... | .................... | .................... |
| A | 2853 hCoV-19/Canada/BC_7515663/2020\|EPI_ISL_418845\|2020 | .................... | .................... | .................... | .................... | .................... | .................... |
| A | 2854 hCoV-19/Canada/BC_7553799/2020\|EPI_ISL_418846\|2020 | .................... | .................... | .................... | .................... | .................... | .................... |
| A | 2855 hCoV-19/Canada/BC_8150537/2020\|EPI_ISL_418847\|2020 | .................... | .................... | .................... | .................... | .................... | .................... |
| A | 2856 hCoV-19/Canada/BC_8159203/2020\|EPI_ISL_418848\|2020 | .................... | .................... | .................... | .................... | .................... | .................... |
| A | 2857 hCoV-19/Canada/BC_8486790/2020\|EPI_ISL_418849\|2020 | .................... | .................... | .................... | .................... | .................... | .................... |
| A | 2858 hCoV-19/Canada/BC_9345042/2020\|EPI_ISL_418856\|2020 | .................... | .................... | .................... | .................... | .................... | .................... |
| A | 2859 hCoV-19/Canada/BC_9345715/2020\|EPI_ISL_418857\|2020 | .................... | .................... | .................... | .................... | .................... | .................... |

```
A  2860 hCoV-19/Hangzhou/ZJU-010/2020|EPI_ISL_418990|2020-  ···················· ···················· ···················· ···················· ···················· ····················
A  2861 hCoV-19/Singapore/20/2020|EPI_ISL_418993|2020-01-2  ···················· ···················· ···················· ···················· ···················· ····················
A  2862 hCoV-19/Singapore/3Clin/2020|EPI_ISL_418994|2020-0  ···················· ···················· ···················· ···················· ···················· ····················
A  2864 hCoV-19/England/20148004904/2020|EPI_ISL_422001|20  ···················· ···················· ···················· ···················· ···················· ····················
A  2865 hCoV-19/England/20148006104/2020|EPI_ISL_422002|20  ···················· ···················· ···················· ···················· ···················· ····················
A  2866 hCoV-19/England/20148006304/2020|EPI_ISL_422003|20  ···················· ···················· ···················· ···················· ···················· ····················
A  2867 hCoV-19/England/20148007004/2020|EPI_ISL_422007|20  ···················· ···················· ···················· ···················· ···················· ····················
A  2869 hCoV-19/England/20148039404/2020|EPI_ISL_422009|20  ···················· ···················· ···················· ···················· ···················· ····················
A  2871 hCoV-19/England/20148046704/2020|EPI_ISL_422011|20  ···················· ···················· ···················· ···················· ···················· ····················
A  2872 hCoV-19/Wales/PHWC-26796/2020|EPI_ISL_422016|2020-  ···················· ···················· ···················· ···················· ···················· ····················
A  2873 hCoV-19/Wales/PHWC-2711F/2020|EPI_ISL_422017|2020-  ···················· ···················· ···················· ···················· ···················· ····················
A  2874 hCoV-19/Wales/PHWC-2673C/2020|EPI_ISL_422018|2020-  ···················· ···················· ···················· ···················· ···················· ····················
A  2875 hCoV-19/Wales/PHWC-265E7/2020|EPI_ISL_422019|2020-  ···················· ···················· ···················· ···················· ···················· ····················
A  2876 hCoV-19/Wales/PHWC-2682A/2020|EPI_ISL_422020|2020-  ···················· ···················· ···················· ···················· ···················· ····················
A  2877 hCoV-19/Wales/PHWC-2665D/2020|EPI_ISL_422021|2020-  ···················· ···A················ ···················· ···················· ···················· ····················
A  2878 hCoV-19/Wales/PHWC-26D49/2020|EPI_ISL_422022|2020-  ···················· ···················· ···················· ···················· ···················· ····················
A  2879 hCoV-19/Wales/PHWC-26981/2020|EPI_ISL_422023|2020-  ···················· ···················· ···················· ···················· ···················· ····················
A  2880 hCoV-19/Wales/PHWC-26B12/2020|EPI_ISL_422024|2020-  ···················· ···················· ···················· ···················· ···················· ····················
A  2881 hCoV-19/Wales/PHWC-263DE/2020|EPI_ISL_422025|2020-  ···················· ···················· ···················· ···················· ···················· ····················
A  2882 hCoV-19/Wales/PHWC-263FC/2020|EPI_ISL_422026|2020-  ···················· ···A················ ···················· ···················· ···················· ····················
A  2883 hCoV-19/Wales/PHWC-2672D/2020|EPI_ISL_422027|2020-  ···················· ···················· ···················· ···················· ···················· ····················
A  2884 hCoV-19/Wales/PHWC-2631A/2020|EPI_ISL_422028|2020-  ···················· ···A················ ···················· ···················· ···················· ····················
A  2885 hCoV-19/Wales/PHWC-25F44/2020|EPI_ISL_422029|2020-  ···················· ···················· ···················· ···················· ···················· ····················
A  2886 hCoV-19/Wales/PHWC-270B8/2020|EPI_ISL_422030|2020-  ···················· ···················· ···················· ···················· ···················· ····················
A  2887 hCoV-19/Wales/PHWC-26DD0/2020|EPI_ISL_422031|2020-  ···················· ···················· ···················· ···················· ···················· ····················
A  2888 hCoV-19/Wales/PHWC-2649F/2020|EPI_ISL_422032|2020-  ···················· ···················· ···················· ···················· ···················· ····················
A  2889 hCoV-19/Wales/PHWC-2681B/2020|EPI_ISL_422033|2020-  ···················· ···················· ···················· ···················· ···················· ····················
A  2890 hCoV-19/Wales/PHWC-27012/2020|EPI_ISL_422034|2020-  ···················· ···················· ···················· ···················· ···················· ····················
A  2891 hCoV-19/Wales/PHWC-26D58/2020|EPI_ISL_422035|2020-  ···················· ···················· ···················· ···················· ···················· ····················
A  2892 hCoV-19/Wales/PHWC-26A15/2020|EPI_ISL_422036|2020-  ···················· ···················· ···················· ···················· ···················· ····················
A  2893 hCoV-19/Wales/PHWC-2663F/2020|EPI_ISL_422037|2020-  ···················· ···················· ···················· ···················· ···················· ····················
A  2894 hCoV-19/Wales/PHWC-2621D/2020|EPI_ISL_422038|2020-  ···················· ···················· ···················· ···················· ···················· ····················
A  2896 hCoV-19/England/20138078204/2020|EPI_ISL_423324|20  ···················· ···················· ···················· ···················· ···················· ····················
A  2897 hCoV-19/England/20138084904/2020|EPI_ISL_423328|20  ···················· ···················· ···················· ···················· ···················· ····················
A  2898 hCoV-19/England/20139002204/2020|EPI_ISL_423337|20  ···················· ···················· ···················· ···················· ···················· ····················
A  2901 hCoV-19/England/20139016704/2020|EPI_ISL_423346|20  ···················· ···················· ···················· ···················· ···················· ····················
A  2904 hCoV-19/England/20139064904/2020|EPI_ISL_423352|20  ···················· ···················· ···················· ···················· ···················· ····················
A  2905 hCoV-19/England/20139065204/2020|EPI_ISL_423353|20  ···················· ···················· ···················· ···················· ···················· ····················
A  2906 hCoV-19/England/20139065404/2020|EPI_ISL_423354|20  ···················· ···················· ···················· ···················· ···················· ····················
A  2907 hCoV-19/England/20139065604/2020|EPI_ISL_423355|20  ···················· ···················· ···················· ···················· ···················· ····················
A  2908 hCoV-19/England/20139067504/2020|EPI_ISL_423356|20  ···················· ···················· ···················· ···················· ···················· ····················
A  2909 hCoV-19/England/20139068504/2020|EPI_ISL_423357|20  ···················· ···················· ···················· ···················· ···················· ····················
A  2910 hCoV-19/England/20139068804/2020|EPI_ISL_423358|20  ···················· ···················· ···················· ···················· ···················· ····················
A  2911 hCoV-19/England/20139069104/2020|EPI_ISL_423359|20  ···················· ···················· ···················· ···················· ···················· ·N··N···············
A  2912 hCoV-19/England/20139069404/2020|EPI_ISL_423360|20  ···················· ···················· ···················· ···················· ···················· ····················
A  2917 hCoV-19/England/20139072204/2020|EPI_ISL_423365|20  ···················· ···················· ···················· ···················· ···················· ····················
A  2918 hCoV-19/England/20140000304/2020|EPI_ISL_423366|20  ···················· ···················· ···················· ···················· ···················· ····················
A  2920 hCoV-19/England/20140000604/2020|EPI_ISL_423368|20  ···················· ···················· ···················· ···················· ···················· ····················
A  2922 hCoV-19/England/20144008304/2020|EPI_ISL_423500|20  ···················· ···················· ···················· ···················· ···················· ····················
A  2923 hCoV-19/England/20144008904/2020|EPI_ISL_423502|20  ···················· ···················· ···················· ···················· ···················· ····················
A  2924 hCoV-19/England/20144009004/2020|EPI_ISL_423503|20  ···················· ···················· ···················· ···················· ············Y······· ····················
A  2925 hCoV-19/England/20144009104/2020|EPI_ISL_423504|20  ···················· ···················· ···················· ···················· ···················· ····················
A  2926 hCoV-19/England/20144011504/2020|EPI_ISL_423506|20  ···················· ···················· ···················· ···················· ···················· ····················
A  2928 hCoV-19/England/20144013704/2020|EPI_ISL_423508|20  ···················· ···················· ···················· ···················· ···················· ····················
A  2929 hCoV-19/England/20144014504/2020|EPI_ISL_423509|20  ···················· ···················· ···················· ···················· ···················· ····················
A  2932 hCoV-19/Iceland/593/2020|EPI_ISL_424613|2020-03-27  ···················· ···················· ···················· ···················· ···················· ····················
A  2933 hCoV-19/Iceland/594/2020|EPI_ISL_424614|2020-03-27  ···················· ···················· ···················· ···················· ···················· ····················
A  2934 hCoV-19/Iceland/595/2020|EPI_ISL_424615|2020-03-27  ···················· ···················· ···················· ···················· ···················· ····················
A  2935 hCoV-19/Iceland/596/2020|EPI_ISL_424616|2020-03-28  ···················· ···················· ···················· ···················· ···················· ····················
A  2936 hCoV-19/Iceland/597/2020|EPI_ISL_424617|2020-03-28  ···················· ···················· ···················· ···················· ···················· ····················
A  2937 hCoV-19/Iceland/599/2020|EPI_ISL_424619|2020-03-28  ···················· ···················· ···················· ···················· ···················· ····················
A  2938 hCoV-19/Iceland/601/2020|EPI_ISL_424621|2020-03-28  ···················· ···················· ···················· ···················· ···················· ····················
A  2939 hCoV-19/Iceland/602/2020|EPI_ISL_424622|2020-03-28  ···················· ···················· ···················· ···················· ···················· ····················
A  2940 hCoV-19/Iceland/604/2020|EPI_ISL_424624|2020-03-28  ···················· ···················· ···················· ···················· ···················· ····················
A  2941 hCoV-19/Mexico/CMX-INER-04/2020|EPI_ISL_424626|202  ···················· ···················· ···················· ···················· ···················· ····················
A  2942 hCoV-19/Mexico/CMX-INER-05/2020|EPI_ISL_424627|202  ···················· ···················· ···················· ···················· ···················· ····················
A  2943 hCoV-19/Belgium/ULG-10066/2020|EPI_ISL_424629|2020  ···················· ···················· ···················· ···················· ···················· ····················
A  2944 hCoV-19/Belgium/ULG-10067/2020|EPI_ISL_424630|2020  ···················· ···················· ···················· ···················· ···················· ····················
A  2945 hCoV-19/Belgium/ULG-10072/2020|EPI_ISL_424631|2020  ···················· ···················· ···················· ···················· ···················· ····················
A  2946 hCoV-19/Belgium/ULG-10069/2020|EPI_ISL_424633|2020  ···················· ···················· ···················· ···················· ···················· ····················
A  2947 hCoV-19/Belgium/ULG-10068/2020|EPI_ISL_424636|2020  ···················· ···················· ···················· ···················· ···················· ····················
A  2948 hCoV-19/Belgium/ULG-10078/2020|EPI_ISL_424637|2020  ···················· ···················· ···················· ···················· ···················· ····················
A  2949 hCoV-19/Belgium/ULG-10081/2020|EPI_ISL_424640|2020  ···················· ···················· ···················· ···················· ···················· ····················
A  2950 hCoV-19/Belgium/ULG-10080/2020|EPI_ISL_424641|2020  ···················· ···················· ···················· ···················· ···················· ····················
A  2951 hCoV-19/Belgium/ULG-10077/2020|EPI_ISL_424642|2020  ···················· ···················· ···················· ···················· ···················· ····················
A  2952 hCoV-19/Belgium/ULG-10089/2020|EPI_ISL_424643|2020  ···················· ···················· ···················· ···················· ···················· ····················
A  2953 hCoV-19/Belgium/ULG-10086/2020|EPI_ISL_424644|2020  ···················· ···················· ···················· ···················· ···················· ····················
A  2954 hCoV-19/Belgium/ULG-10085/2020|EPI_ISL_424645|2020  ···················· ···················· ···················· ···················· ···················· ····················
```

| | | | | | | | | |
|---|---|---|---|---|---|---|---|---|
| A | 2955 hCoV-19/Belgium/ULG-10088/2020\|EPI_ISL_424646\|2020 | ···················· | ···················· | ···················· | ···················· | ···················· | ···················· | |
| A | 2956 hCoV-19/Belgium/ULG-10092/2020\|EPI_ISL_424647\|2020 | ···················· | ···················· | ···················· | ···················· | ···················· | ···················· | |
| A | 2957 hCoV-19/Belgium/ULG-10082/2020\|EPI_ISL_424649\|2020 | ···················· | ···················· | ···················· | ···················· | ···················· | ···················· | |
| A | 2958 hCoV-19/Belgium/ULG-10094/2020\|EPI_ISL_424650\|2020 | ···················· | ···················· | ···················· | ···················· | ···················· | ···················· | |
| A | 2959 hCoV-19/Belgium/ULG-10095/2020\|EPI_ISL_424651\|2020 | ···················· | ···················· | ···················· | ···················· | ···················· | ···················· | |
| A | 2960 hCoV-19/Belgium/ULG-10055/2020\|EPI_ISL_424652\|2020 | ···················· | ···················· | ···················· | ···················· | ···················· | ···················· | |
| A | 2961 hCoV-19/Belgium/ULG-10058/2020\|EPI_ISL_424653\|2020 | ···················· | ···················· | ···················· | ···················· | ···················· | ···················· | |
| A | 2962 hCoV-19/Belgium/ULG-10057/2020\|EPI_ISL_424654\|2020 | ···················· | ···················· | ···················· | ···················· | ···················· | ···················· | |
| A | 2963 hCoV-19/Belgium/ULG-10056/2020\|EPI_ISL_424655\|2020 | ···················· | ···················· | ···················· | ···················· | ···················· | ···················· | |
| A | 2964 hCoV-19/Belgium/ULG-10052/2020\|EPI_ISL_424657\|2020 | ···················· | ···················· | ···················· | ···················· | ···················· | ···················· | |
| A | 2965 hCoV-19/Belgium/ULG-10049/2020\|EPI_ISL_424658\|2020 | ···················· | ···················· | ···················· | ···················· | ···················· | ···················· | |
| A | 2966 hCoV-19/Belgium/ULG-10051/2020\|EPI_ISL_424660\|2020 | ···················· | ···················· | ···················· | ···················· | ···················· | ···················· | |
| A | 2967 hCoV-19/Belgium/ULG-10053/2020\|EPI_ISL_424661\|2020 | ···················· | ···················· | ···················· | ···················· | ···················· | ···················· | |
| A | 2968 hCoV-19/Belgium/ULG-10061/2020\|EPI_ISL_424662\|2020 | ···················· | ··········A········· | ···················· | ···················· | ···················· | ···················· | |
| A | 2969 hCoV-19/Belgium/ULG-10062/2020\|EPI_ISL_424664\|2020 | ···················· | ···················· | ···················· | ···················· | ···················· | ···················· | |
| A | 2970 hCoV-19/Mexico/CMX-InDRE_03/2020\|EPI_ISL_424667\|20 | ···················· | ···················· | ···················· | ···················· | ···················· | ···················· | |
| A | 2971 hCoV-19/USA/AZ-ASU2922/2020\|EPI_ISL_424668\|2020-03 | ···················· | ···················· | ···················· | ···················· | ···················· | ···················· | |
| A | 2972 hCoV-19/USA/AZ-ASU2923/2020\|EPI_ISL_424669\|2020-03 | ···················· | ···················· | ···················· | ···················· | ···················· | ···················· | |
| A | 2973 hCoV-19/Mexico/QUE-InDRE-04/2020\|EPI_ISL_424670\|20 | ···················· | ···················· | ···················· | ···················· | ···················· | ···················· | |
| A | 2974 hCoV-19/USA/AZ-ASU2936/2020\|EPI_ISL_424671\|2020-03 | ···················· | ···················· | ···················· | ···················· | ···················· | ···················· | |
| A | 2975 hCoV-19/Mexico/PUE-InDRE-05/2020\|EPI_ISL_424672\|20 | ···················· | ···················· | ···················· | ···················· | ···················· | ···················· | |
| A | 2976 hCoV-19/Mexico/CMX-InDRE-06/2020\|EPI_ISL_424673\|20 | ···················· | ···················· | ···················· | ···················· | ···················· | ···················· | |
| A | 2977 hCoV-19/USA/SC_3569/2020\|EPI_ISL_424841\|2020-03-06 | ···················· | ···················· | ···················· | ···················· | ···················· | ···················· | |
| A | 2978 hCoV-19/USA/SC_3575/2020\|EPI_ISL_424842\|2020-03-07 | ···················· | ···················· | ···················· | ···················· | ···················· | ···················· | |
| A | 2979 hCoV-19/USA/MA_0020/2020\|EPI_ISL_424843\|2020-03-05 | ···················· | ···················· | ···················· | ···················· | ···················· | ···················· | |
| A | 2980 hCoV-19/USA/MA_3672/2020\|EPI_ISL_424844\|2020-03-06 | ···················· | ···················· | ···················· | ···················· | ···················· | ···················· | |
| A | 2981 hCoV-19/USA/MA_3699/2020\|EPI_ISL_424845\|2020-03-06 | ···················· | ···················· | ···················· | ···················· | ···················· | ···················· | |
| A | 2982 hCoV-19/USA/MA_3878/2020\|EPI_ISL_424846\|2020-03-06 | ···················· | ···················· | ···················· | ···················· | ···················· | ···················· | |
| A | 2983 hCoV-19/USA/MA_4811/2020\|EPI_ISL_424847\|2020-03-07 | ···················· | ···················· | ···················· | ···················· | ···················· | ···················· | |
| A | 2984 hCoV-19/USA/AZ_8132/2020\|EPI_ISL_424848\|2020-03-09 | ···················· | ···················· | ···················· | ···················· | ···················· | ···················· | |
| A | 2985 hCoV-19/USA/AZ_8135/2020\|EPI_ISL_424849\|2020-03-09 | ···················· | ···················· | ···················· | ···················· | ···················· | ···················· | |
| A | 2986 hCoV-19/USA/IL_0087/2020\|EPI_ISL_424850\|2020-03-07 | ···················· | ···················· | ···················· | ···················· | ···················· | ···················· | |
| A | 2987 hCoV-19/USA/IL_0089/2020\|EPI_ISL_424851\|2020-03-07 | ···················· | ···················· | ···················· | ···················· | ···················· | ···················· | |
| A | 2988 hCoV-19/USA/DC_0004/2020\|EPI_ISL_424852\|2020-03-06 | ···················· | ···················· | ···················· | ···················· | ···················· | ···················· | |
| A | 2989 hCoV-19/USA/FL_9590/2020\|EPI_ISL_424853\|2020-03-05 | ···················· | ···················· | ···················· | ···················· | ···················· | ···················· | |
| A | 2990 hCoV-19/USA/FL_9655/2020\|EPI_ISL_424854\|2020-03-05 | ···················· | ···················· | ···················· | ···················· | ···················· | ···················· | |
| A | 2991 hCoV-19/Scotland/EDB214/2020\|EPI_ISL_425944\|2020-0 | ···················· | ···················· | ···················· | ···················· | ···················· | ···················· | |
| A | 2992 hCoV-19/Scotland/EDB216/2020\|EPI_ISL_425945\|2020-0 | ···················· | ···················· | ···················· | ···················· | ···················· | ···················· | |
| A | 2993 hCoV-19/Scotland/EDB218/2020\|EPI_ISL_425946\|2020-0 | ···················· | ···················· | ···················· | ···················· | ···················· | ···················· | |
| A | 2994 hCoV-19/Scotland/EDB219/2020\|EPI_ISL_425947\|2020-0 | ···················· | ···················· | ···················· | ···················· | ···················· | ···················· | |
| A | 2995 hCoV-19/Scotland/EDB222/2020\|EPI_ISL_425948\|2020-0 | ···················· | ···················· | ···················· | ···················· | ···················· | ···················· | |
| A | 2996 hCoV-19/Scotland/EDB226/2020\|EPI_ISL_425949\|2020-0 | ···················· | ···················· | ···················· | ···················· | ···················· | ···················· | |
| A | 2997 hCoV-19/Scotland/EDB227/2020\|EPI_ISL_425950\|2020-0 | ···················· | ···················· | ···················· | ···················· | ···················· | ···················· | |
| A | 2998 hCoV-19/Scotland/EDB232/2020\|EPI_ISL_425951\|2020-0 | ···················· | ···················· | ···················· | ···················· | ···················· | ···················· | |
| A | 2999 hCoV-19/Scotland/EDB234/2020\|EPI_ISL_425952\|2020-0 | ···················· | ···················· | ···················· | ···················· | ···················· | ···················· | |
| A | 3000 hCoV-19/Scotland/EDB245/2020\|EPI_ISL_425953\|2020-0 | ···················· | ···················· | ···················· | ···················· | ···················· | ···················· | |
| A | 3001 hCoV-19/Scotland/EDB248/2020\|EPI_ISL_425954\|2020-0 | ···················· | ···················· | ···················· | ···················· | ···················· | ···················· | |
| A | 3002 hCoV-19/Scotland/EDB252/2020\|EPI_ISL_425955\|2020-0 | ···················· | ···················· | ···················· | ···················· | ···················· | ···················· | |
| A | 3003 hCoV-19/Scotland/EDB254/2020\|EPI_ISL_425956\|2020-0 | ···················· | ···················· | ···················· | ···················· | ···················· | ···················· | |
| A | 3004 hCoV-19/Scotland/EDB270/2020\|EPI_ISL_425958\|2020-0 | ···················· | ···················· | ···················· | ···················· | ···················· | ···················· | |
| A | 3005 hCoV-19/Scotland/EDB273/2020\|EPI_ISL_425959\|2020-0 | ···················· | ···················· | ···················· | ···················· | ···················· | ···················· | |
| A | 3006 hCoV-19/Scotland/EDB275/2020\|EPI_ISL_425961\|2020-0 | ···················· | ···················· | ···················· | ···················· | ···················· | ···················· | |
| A | 3007 hCoV-19/Scotland/EDB278/2020\|EPI_ISL_425962\|2020-0 | ···················· | ···················· | ···················· | ···················· | ···················· | ···················· | |
| A | 3008 hCoV-19/Scotland/EDB283/2020\|EPI_ISL_425963\|2020-0 | ···················· | ···················· | ···················· | ···················· | ···················· | ···················· | |
| A | 3009 hCoV-19/Scotland/EDB287/2020\|EPI_ISL_425964\|2020-0 | ···················· | ···················· | ···················· | ···················· | ···················· | ···················· | |
| A | 3010 hCoV-19/Scotland/EDB288/2020\|EPI_ISL_425965\|2020-0 | ···················· | ···················· | ···················· | ···················· | ···················· | ···················· | |
| A | 3011 hCoV-19/Scotland/EDB297/2020\|EPI_ISL_425966\|2020-0 | ···················· | ···················· | ···················· | ···················· | ···················· | ···················· | |
| A | 3012 hCoV-19/Scotland/EDB300/2020\|EPI_ISL_425967\|2020-0 | ···················· | ···················· | ···················· | ···················· | ···················· | ···················· | |
| A | 3013 hCoV-19/Scotland/EDB314/2020\|EPI_ISL_425968\|2020-0 | ···················· | ···················· | ···················· | ···················· | ···················· | ···················· | |
| A | 3014 hCoV-19/Scotland/EDB318/2020\|EPI_ISL_425969\|2020-0 | ···················· | ···················· | ···················· | ···················· | ···················· | ···················· | |
| A | 3015 hCoV-19/Scotland/EDB325/2020\|EPI_ISL_425971\|2020-0 | ···················· | ···················· | ···················· | ···················· | ···················· | ···················· | |
| A | 3016 hCoV-19/Scotland/EDB326/2020\|EPI_ISL_425972\|2020-0 | ···················· | ···················· | ···················· | ···················· | ···················· | ···················· | |
| A | 3019 hCoV-19/Scotland/EDB342/2020\|EPI_ISL_425976\|2020-0 | ···················· | ···················· | ···················· | ···················· | ···················· | ···················· | |
| A | 3020 hCoV-19/Scotland/EDB354/2020\|EPI_ISL_425978\|2020-0 | ···················· | ···················· | ···················· | ···················· | ···················· | ···················· | |
| A | 3021 hCoV-19/Scotland/EDB355/2020\|EPI_ISL_425979\|2020-0 | ···················· | ···················· | ···················· | ···················· | ···················· | ···················· | |
| A | 3022 hCoV-19/Scotland/EDB356/2020\|EPI_ISL_425980\|2020-0 | ···················· | ···················· | ···················· | ···················· | ···················· | ···················· | |
| A | 3023 hCoV-19/Scotland/EDB363/2020\|EPI_ISL_425982\|2020-0 | ···················· | ···················A | ···················· | ···················· | ···················· | ···················· | 1 |
| A | 3025 hCoV-19/Scotland/EDB373/2020\|EPI_ISL_425985\|2020-0 | ···················· | ···················· | ···················· | ···················· | ···················· | ···················· | |
| A | 3026 hCoV-19/Scotland/EDB374/2020\|EPI_ISL_425986\|2020-0 | ···················· | ···················· | ···················· | ···················· | ···················· | ···················· | |
| A | 3027 hCoV-19/Scotland/EDB378/2020\|EPI_ISL_425987\|2020-0 | ···················· | ···················· | ···················· | ···················· | ···················· | ···················· | |
| A | 3028 hCoV-19/Scotland/EDB379/2020\|EPI_ISL_425988\|2020-0 | ···················· | ···················· | ···················· | ···················· | ···················· | ···················· | |
| A | 3029 hCoV-19/Scotland/EDB383/2020\|EPI_ISL_425990\|2020-0 | ···················· | ···················· | ···················· | ···················· | ···················· | ···················· | |
| A | 3030 hCoV-19/Scotland/EDB386/2020\|EPI_ISL_425991\|2020-0 | ···················· | ···················· | ···················· | ···················· | ···················· | ···················· | |
| A | 3031 hCoV-19/Scotland/EDB389/2020\|EPI_ISL_425992\|2020-0 | ···················· | ···················· | ···················· | ···················· | ···················· | ···················· | |
| A | 3032 hCoV-19/Scotland/EDB390/2020\|EPI_ISL_425993\|2020-0 | ···················· | ···················· | ···················· | ···················· | ···················· | ···················· | |
| A | 3033 hCoV-19/Scotland/EDB391/2020\|EPI_ISL_425994\|2020-0 | ···················· | ···················· | ···················· | ···················· | ···················· | ···················· | |
| A | 3034 hCoV-19/Scotland/EDB392/2020\|EPI_ISL_425995\|2020-0 | ···················· | ···················· | ···················· | ···················· | ···················· | ···················· | |
| A | 3035 hCoV-19/Scotland/EDB396/2020\|EPI_ISL_425996\|2020-0 | ··················T· | ···················· | ···················· | ···················· | ···················· | ···················· | 1 |

```
A   3036 hCoV-19/Scotland/EDB397/2020|EPI_ISL_425997|2020-0   ···················· ···················· ····················· ···················· ···················· ····················
A   3037 hCoV-19/Scotland/EDB399/2020|EPI_ISL_425998|2020-0   ···················· ···················· ····················· ···················· ···················· ····················
A   3038 hCoV-19/Scotland/EDB401/2020|EPI_ISL_425999|2020-0   ···················· ···················· ····················· ···················· ···················· ····················
A   3039 hCoV-19/Australia/VIC779/2020|EPI_ISL_427054|2020-   ···················· ···················· ····················· ···················· ···················· ····················
A   3040 hCoV-19/Australia/VIC780/2020|EPI_ISL_427055|2020-   ···················· ···················· ····················· ···················· ···················· ····················
A   3044 hCoV-19/Australia/VIC785/2020|EPI_ISL_427060|2020-   ···················· ···················· ····················· ···················· ···················· ····················
A   3061 hCoV-19/Australia/VIC807/2020|EPI_ISL_427081|2020-   ···················· ···················· ····················· ···················· ···················· ····················
A   3062 hCoV-19/Australia/VIC808/2020|EPI_ISL_427082|2020-   ···················· ···················· ····················· ···················· ···················· ····················
A   3063 hCoV-19/Australia/VIC812/2020|EPI_ISL_427085|2020-   ···················· ···················· ····················· ···················· ···················· ····················
A   3064 hCoV-19/Australia/VIC820/2020|EPI_ISL_427092|2020-   ···················· ···················· ····················· ···················· ···················· ····················
A   3065 hCoV-19/Australia/VIC821/2020|EPI_ISL_427093|2020-   ···················· ···················· ····················· ···················· ···················· ····················
A   3066 hCoV-19/Australia/VIC823/2020|EPI_ISL_427094|2020-   ···················· ···················· ····················· ···················· ···················· ····················
A   3067 hCoV-19/Australia/VIC824/2020|EPI_ISL_427095|2020-   ···················· ···················· ····················· ···················· ···················· ····················
A   3068 hCoV-19/Australia/VIC828/2020|EPI_ISL_427096|2020-   ···················· ···················· ····················· ···················· ···················· ····················
A   3069 hCoV-19/USA/CT-Yale-076/2020|EPI_ISL_428384|2020-0   ···················· ···················· ····················· ···················· ···················· ····················
A   3070 hCoV-19/USA/CT-Yale-066/2020|EPI_ISL_428385|2020-0   ···················· ···················· ····················· ···················· ···················· ····················
A   3071 hCoV-19/USA/CT-Yale-062/2020|EPI_ISL_428387|2020-0   ···················· ···················· ····················· ···················· ···················· ····················
A   3072 hCoV-19/USA/CT-Yale-060/2020|EPI_ISL_428391|2020-0   ···················· ···················· ····················· ···················· ··············N····· ····················
A   3073 hCoV-19/Wales/PHW04/2020|EPI_ISL_415041|2020-03-07   ···················· ···················· ····················· ···················· ···················· ····················
A   3074 hCoV-19/Shanghai/SH0005/2020|EPI_ISL_416319|2020-0   ···················· ···················· ····················· ···················· ···················· ····················
A   3075 hCoV-19/Shanghai/SH0007/2020|EPI_ISL_416320|2020-0   ···················· ···················· ····················· ···················· ···················· ····················
A   3076 hCoV-19/Shanghai/SH0008/2020|EPI_ISL_416321|2020-0   ···················· ···················· ····················· ···················· ···················· ····················
A   3077 hCoV-19/Shanghai/SH0009/2020|EPI_ISL_416322|2020-0   ···················· ···················· ····················· ···················· ···················· ····················
A   3078 hCoV-19/Shanghai/SH0010/2020|EPI_ISL_416323|2020-0   ···················· ···················· ····················· ···················· ···················· ····················
A   3079 hCoV-19/Shanghai/SH0011/2020|EPI_ISL_416324|2020-0   ···················· ···················· ····················· ···················· ···················· ····················
A   3080 hCoV-19/Shanghai/SH0012/2020|EPI_ISL_416325|2020-0   ···················· ···················· ····················· ···················· ···················· ····················
A   3081 hCoV-19/Shanghai/SH0013/2020|EPI_ISL_416326|2020-0   ···················· ···················· ····················· ···················· ···················· ····················
A   3082 hCoV-19/Shanghai/SH0020/2020|EPI_ISL_416329|2020-0   ···················· ···················· ····················· ···················· ···················· ····················
A   3083 hCoV-19/Shanghai/SH0021/2020|EPI_ISL_416330|2020-0   ···················· ···················· ····················· ···················· ···················· ····················
A   3084 hCoV-19/Shanghai/SH0023/2020|EPI_ISL_416332|2020-0   ···················· ···················· ····················· ···················· ···················· ····················
A   3085 hCoV-19/Shanghai/SH0024/2020|EPI_ISL_416333|2020-0   ···················· ···················· ····················· ···················· ···················· ····················
A   3086 hCoV-19/Shanghai/SH0025/2020|EPI_ISL_416334|2020-0   ···················· ···················· ····················· ···················· ···················· ····················
A   3087 hCoV-19/Shanghai/SH0026/2020|EPI_ISL_416335|2020-0   ···················· ···················· ····················· ···················· ···················· ····················
A   3088 hCoV-19/Shanghai/SH0027/2020|EPI_ISL_416336|2020-0   ···················· ···················· ····················· ···················· ···················· ····················
A   3089 hCoV-19/Shanghai/SH0028/2020|EPI_ISL_416337|2020-0   ···················· ···················· ····················· ···················· ···················· ····················
A   3090 hCoV-19/Shanghai/SH0029/2020|EPI_ISL_416338|2020-0   ···················· ···················· ····················· ···················· ···················· ····················
A   3091 hCoV-19/Shanghai/SH0030/2020|EPI_ISL_416339|2020-0   ···················· ···················· ····················· ···················· ···················· ····················
A   3092 hCoV-19/Shanghai/SH0031/2020|EPI_ISL_416340|2020-0   ···················· ···················· ····················· ···················· ···················· ····················
A   3093 hCoV-19/Shanghai/SH0032/2020|EPI_ISL_416341|2020-0   ···················· ···················· ····················· ···················· ···················· ····················
A   3094 hCoV-19/Shanghai/SH0033/2020|EPI_ISL_416342|2020-0   ···················· ···················· ····················· ···················· ···················· ····················
A   3095 hCoV-19/Shanghai/SH0039/2020|EPI_ISL_416348|2020-0   ···················· ···················· ····················· ···················· ···················· ····················
A   3096 hCoV-19/Shanghai/SH0040/2020|EPI_ISL_416349|2020-0   ···················· ···················· ····················· ···················· ···················· ····················
A   3098 hCoV-19/Shanghai/SH0043/2020|EPI_ISL_416352|2020-0   ···················· ···················· ····················· ···················· ···················· ····················
A   3099 hCoV-19/Shanghai/SH0044/2020|EPI_ISL_416353|2020-0   ···················· ···················· ····················· ···················· ···················· ····················
A   3100 hCoV-19/Shanghai/SH0045/2020|EPI_ISL_416354|2020-0   ···················· ···················· ····················· ···················· ···················· ····················
A   3101 hCoV-19/Shanghai/SH0047/2020|EPI_ISL_416355|2020-0   ···················· ···················· ····················· ···················· ···················· ····················
A   3102 hCoV-19/Shanghai/SH0050/2020|EPI_ISL_416358|2020-0   ···················· ·A·················· ····················· ···················· ···················· ····················
A   3103 hCoV-19/Shanghai/SH0051/2020|EPI_ISL_416359|2020-0   ···················· ···················· ····················· ···················· ···················· ····················
A   3104 hCoV-19/Shanghai/SH0053/2020|EPI_ISL_416361|2020-0   ···················· ···················· ····················· ···················· ···················· ····················
A   3105 hCoV-19/Shanghai/SH0054/2020|EPI_ISL_416362|2020-0   ···················· ··········A········· ····················· ···················· ···················· ····················
A   3106 hCoV-19/Shanghai/SH0055/2020|EPI_ISL_416363|2020-0   ···················· ···················· ····················· ···················· ···················· ····················
A   3107 hCoV-19/Shanghai/SH0056/2020|EPI_ISL_416364|2020-0   ···················· ···················· ····················· ···················· ···················· ····················
A   3108 hCoV-19/Shanghai/SH0058/2020|EPI_ISL_416365|2020-0   ···················· ·A·················· ····················· ···················· ···················· ····················
A   3109 hCoV-19/Shanghai/SH0059/2020|EPI_ISL_416366|2020-0   ···················· ···················· ····················· ···················· ···················· ····················
A   3110 hCoV-19/Shanghai/SH0060/2020|EPI_ISL_416367|2020-0   ···················· ···················· ····················· ···················· ···················· ····················
A   3111 hCoV-19/Shanghai/SH0065/2020|EPI_ISL_416368|2020-0   ···················· ···················· ····················· ···················· ···················· ····NN··············
A   3112 hCoV-19/Shanghai/SH0066/2020|EPI_ISL_416369|2020-0   ···················· ···················· ····················· ···················· ···················· ····················
A   3113 hCoV-19/Shanghai/SH0067/2020|EPI_ISL_416370|2020-0   ···················· ···················· ····················· ···················· ···················· ····················
A   3114 hCoV-19/Shanghai/SH0069/2020|EPI_ISL_416372|2020-0   ···················· ···················· ····················· ···················· ···················· ····················
A   3115 hCoV-19/Shanghai/SH0070/2020|EPI_ISL_416373|2020-0   ···················· ···················· ····················· ···················· ···················· ····················
A   3116 hCoV-19/Shanghai/SH0073/2020|EPI_ISL_416376|2020-0   ···················· ···················· ····················· ···················· ···················· ····················
A   3117 hCoV-19/Shanghai/SH0074/2020|EPI_ISL_416377|2020-0   ···················· ···················· ····················· ···················· ···················· ····················
A   3118 hCoV-19/Shanghai/SH0075/2020|EPI_ISL_416378|2020-0   ···················· ···················· ····················· ···················· ···················· N···················
A   3119 hCoV-19/Shanghai/SH0076/2020|EPI_ISL_416379|2020-0   ···················· ···················· ····················· ···················· ···················· ····················
A   3120 hCoV-19/Shanghai/SH0077/2020|EPI_ISL_416380|2020-0   ···················· ···················· ····················· ···················· ···················· ····················
A   3121 hCoV-19/Shanghai/SH0079/2020|EPI_ISL_416381|2020-0   ···················· ···················· ····················· ···················· ···················· ····················
A   3122 hCoV-19/Shanghai/SH0080/2020|EPI_ISL_416382|2020-0   ···················· ···················· ····················· ···················· ···················· ····················
A   3123 hCoV-19/Shanghai/SH0083/2020|EPI_ISL_416384|2020-0   ···················· ···················· ····················· ···················· ···················· ····················
A   3124 hCoV-19/Shanghai/SH0088/2020|EPI_ISL_416387|2020-0   ···················· ···················· ····················· ···················· ···················· ····················
A   3125 hCoV-19/Shanghai/SH0093/2020|EPI_ISL_416389|2020-0   ···················· ····A··············· ····················· ···················· ···················· ····················
A   3126 hCoV-19/Shanghai/SH0094/2020|EPI_ISL_416390|2020-0   ···················· ···················· ····················· ···················· ···················· ····················
A   3127 hCoV-19/Shanghai/SH0101/2020|EPI_ISL_416393|2020-0   ···················· ···················· ····················· ···················· ···················· ····················
A   3128 hCoV-19/Shanghai/SH0102/2020|EPI_ISL_416394|2020-0   ···················· ···················· ····················· ···················· ···················· ····················
A   3129 hCoV-19/Shanghai/SH0106/2020|EPI_ISL_416396|2020-0   ···················· ···················· ····················· ···················· ···················· ····················
A   3130 hCoV-19/Shanghai/SH0107/2020|EPI_ISL_416397|2020-0   ···················· ···················· ····················· ···················· ···················· ····················
A   3131 hCoV-19/Shanghai/SH0109/2020|EPI_ISL_416398|2020-0   ···················· ···················· ····················· ···················· ···················· ····················
A   3132 hCoV-19/Shanghai/SH0110/2020|EPI_ISL_416399|2020-0   ···················· ···················· ····················· ···················· ···················· ····················
A   3133 hCoV-19/Iceland/321/2020|EPI_ISL_417650|2020-03-18    ···················· ···················· ····················· ···················· ···················· ····················
```

| | | | | | | | |
|---|---|---|---|---|---|---|---|
| A | 3134 hCoV-19/Iceland/322/2020\|EPI_ISL_417651\|2020-03-18 | ···················· | ···················· | ···················· | ···················· | ···················· | ···················· |
| A | 3135 hCoV-19/Iceland/323/2020\|EPI_ISL_417652\|2020-03-18 | ···················· | ···················· | ···················· | ···················· | ···················· | ···················· |
| A | 3136 hCoV-19/Iceland/324/2020\|EPI_ISL_417653\|2020-03-18 | ···················· | ···················· | ···················· | ···················· | ···················· | ···················· |
| A | 3137 hCoV-19/Iceland/325/2020\|EPI_ISL_417654\|2020-03-18 | ···················· | ···················· | ···················· | ···················· | ···················· | ···················· |
| A | 3138 hCoV-19/Iceland/326/2020\|EPI_ISL_417655\|2020-03-14 | ···················· | ···················· | ···················· | ···················· | ···················· | ···················· |
| A | 3140 hCoV-19/Iceland/330/2020\|EPI_ISL_417659\|2020-03-14 | ···················· | ···················· | ···················· | ···················· | ···················· | ···················· |
| A | 3142 hCoV-19/Iceland/335/2020\|EPI_ISL_417664\|2020-03-13 | ···················· | ···················· | ···················· | ···················· | ···················· | ···················· |
| A | 3143 hCoV-19/Iceland/336/2020\|EPI_ISL_417665\|2020-03-14 | ···················· | ···················· | ···················· | ···················· | ···················· | ···················· |
| A | 3144 hCoV-19/Iceland/337/2020\|EPI_ISL_417666\|2020-03-15 | ···················· | ···················· | ···················· | ···················· | ···················· | ···················· |
| A | 3145 hCoV-19/Iceland/338/2020\|EPI_ISL_417667\|2020-03-15 | ···················· | ···················· | ···················· | ···················· | ···················· | ···················· |
| A | 3146 hCoV-19/Iceland/339/2020\|EPI_ISL_417668\|2020-03-15 | ···················· | ···················· | ···················· | ···················· | ···················· | ···················· |
| A | 3147 hCoV-19/Iceland/341/2020\|EPI_ISL_417670\|2020-03-15 | ···················· | ···················· | ···················· | ···················· | ··········N········· | ···················· |
| A | 3148 hCoV-19/Iceland/38/2020\|EPI_ISL_417672\|2020-03-16 | ···················· | ···················· | ···················· | ···················· | ···················· | ···················· |
| A | 3150 hCoV-19/Iceland/98/2020\|EPI_ISL_417676\|2020-03-16 | ···················· | ···················· | ···················· | ···················· | ···················· | ···················· |
| A | 3151 hCoV-19/Iceland/39/2020\|EPI_ISL_417677\|2020-03-04 | ···················· | ···················· | ···················· | ···················· | ···················· | ···················· |
| A | 3152 hCoV-19/Iceland/185/2020\|EPI_ISL_417678\|2020-03-16 | ···················· | ···················· | ···················· | ···················· | ···················· | ···················· |
| A | 3154 hCoV-19/Iceland/177/2020\|EPI_ISL_417680\|2020-03-15 | ···················· | ···················· | ···················· | ···················· | ···················· | ···················· |
| A | 3155 hCoV-19/Iceland/37/2020\|EPI_ISL_417681\|2020-03-04 | ···················· | ···················· | ···················· | ···················· | ···················· | ···················· |
| A | 3156 hCoV-19/Iceland/101/2020\|EPI_ISL_417682\|2020-03-10 | ···················· | ···················· | ···················· | ···················· | ···················· | ···················· |
| A | 3157 hCoV-19/Iceland/47/2020\|EPI_ISL_417683\|2020-03-05 | ···················· | ···················· | ···················· | ···················· | ···················· | ···················· |
| A | 3158 hCoV-19/Iceland/54/2020\|EPI_ISL_417684\|2020-03-06 | ···················· | ···················· | ···················· | ···················· | ···················· | ···················· |
| A | 3159 hCoV-19/Iceland/215/2020\|EPI_ISL_417685\|2020-03-16 | ···················· | ···················· | ···················· | ···················· | ···················· | ···················· |
| A | 3160 hCoV-19/Iceland/55/2020\|EPI_ISL_417687\|2020-03-06 | ···················· | ···················· | ···················· | ···················· | ···················· | ···················· |
| A | 3161 hCoV-19/Iceland/12/2020\|EPI_ISL_417688\|2020-03-01 | ···················· | ···················· | ···················· | ···················· | ···················· | ···················· |
| A | 3162 hCoV-19/Iceland/160/2020\|EPI_ISL_417689\|2020-03-13 | ···················· | ···················· | ···················· | ···················· | ···················· | ···················· |
| A | 3163 hCoV-19/Iceland/125/2020\|EPI_ISL_417690\|2020-03-11 | ···················· | ···················· | ···················· | ···················· | ···················· | ···················· |
| A | 3164 hCoV-19/Iceland/111/2020\|EPI_ISL_417691\|2020-03-11 | ···················· | ···················· | ···················· | ···················· | ···················· | ···················· |
| A | 3165 hCoV-19/Iceland/57/2020\|EPI_ISL_417692\|2020-03-06 | ···················· | ···················· | ···················· | ···················· | ···················· | ···················· |
| A | 3166 hCoV-19/Iceland/25/2020\|EPI_ISL_417693\|2020-03-02 | ···················· | ···················· | ···················· | ···················· | ···················· | ···················· |
| A | 3167 hCoV-19/Iceland/3/2020\|EPI_ISL_417694\|2020-03-13 | ···················· | ···················· | ···················· | ···················· | ···················· | ···················· |
| A | 3168 hCoV-19/Iceland/20/2020\|EPI_ISL_417695\|2020-03-02 | ···················· | ···················· | ···················· | ···················· | ···················· | ···················· |
| A | 3169 hCoV-19/Iceland/135/2020\|EPI_ISL_417696\|2020-03-12 | ···················· | ···················· | ···················· | ···················· | ···················· | ···················· |
| A | 3171 hCoV-19/Iceland/103/2020\|EPI_ISL_417698\|2020-03-10 | ···················· | ···················· | ···················· | ···················· | ···················· | ···················· |
| A | 3172 hCoV-19/Iceland/206/2020\|EPI_ISL_417699\|2020-03-16 | ···················· | ···················· | ···················· | ···················· | ···················· | ···················· |
| A | 3173 hCoV-19/USA/NY-NYUMC34/2020\|EPI_ISL_418980\|2020-03 | ···················· | ···················· | ···················· | ···················· | ···················· | ···················· |
| A | 3174 hCoV-19/Belgium/VRAR-030643/2020\|EPI_ISL_418981\|20 | ···················· | ···················· | ···················· | ···················· | ···················· | ···················· |
| A | 3175 hCoV-19/Belgium/HAA-030953/2020\|EPI_ISL_418982\|202 | ···················· | ···················· | ···················· | ···················· | ···················· | ···················· |
| A | 3176 hCoV-19/Belgium/DWAEE-030954/2020\|EPI_ISL_418983\|2 | ···················· | ···················· | ···················· | ···················· | ···················· | ···················· |
| A | 3177 hCoV-19/Belgium/BG-030955/2020\|EPI_ISL_418984\|2020 | ···················· | ···················· | ···················· | ···················· | ···················· | ···················· |
| A | 3178 hCoV-19/Belgium/LT-030956/2020\|EPI_ISL_418985\|2020 | ···················· | ···················· | ···················· | ···················· | ···················· | ···················· |
| A | 3179 hCoV-19/Belgium/RS-030257/2020\|EPI_ISL_418986\|2020 | ···················· | ···················· | ···················· | ···················· | ···················· | ···················· |
| A | 3180 hCoV-19/Belgium/CG-030158/2020\|EPI_ISL_418987\|2020 | ···················· | ···················· | ···················· | ···················· | ···················· | ···················· |
| A | 3181 hCoV-19/Belgium/JRH-030459/2020\|EPI_ISL_418988\|202 | ···················· | ···················· | ···················· | ···················· | ···················· | ···················· |
| A | 3182 hCoV-19/Belgium/RT-030460/2020\|EPI_ISL_418989\|2020 | ···················· | ···················· | ···················· | ···················· | ···················· | ···················· |
| A | 3183 hCoV-19/Hangzhou/ZJU-011/2020\|EPI_ISL_418991\|2020- | ···················· | ···················· | ···················· | ···················· | ···················· | ···················· |
| A | 3184 hCoV-19/Singapore/17/2020\|EPI_ISL_418992\|2020-02-1 | ···················· | ···················· | ···················· | ···················· | ···················· | ···················· |
| A | 3185 hCoV-19/Singapore/12Clin/2020\|EPI_ISL_418995\|2020- | ···················· | ···················· | ···················· | ···················· | ···················· | ···················· |
| A | 3186 hCoV-19/Singapore/15/2020\|EPI_ISL_418996\|2020-01-2 | ···················· | ···················· | ···················· | ···················· | ···················· | ···················· |
| A | 3187 hCoV-19/Singapore/16/2020\|EPI_ISL_418997\|2020-02-0 | ···················· | ···················· | ···················· | ···················· | ···················· | ···················· |
| A | 3188 hCoV-19/Singapore/14Clin/2020\|EPI_ISL_418998\|2020- | ···················· | ···················· | ···················· | ···················· | ···················· | ···················· |
| A | 3189 hCoV-19/Singapore/18/2020\|EPI_ISL_418999\|2020-03-0 | ···················· | ···················· | ···················· | ···················· | ···················· | ···················· |
| A | 3190 hCoV-19/Wales/PHWC-269CD/2020\|EPI_ISL_422176\|2020- | ···················· | ···················· | ···················· | ···················· | ···················· | ···················· |
| A | 3191 hCoV-19/Wales/PHWC-2668A/2020\|EPI_ISL_422177\|2020- | ···················· | ····A··············· | ···················· | ···················· | ···················· | ···················· |
| A | 3192 hCoV-19/Wales/PHWC-26D67/2020\|EPI_ISL_422178\|2020- | ···················· | ···················· | ···················· | ···················· | ···················· | ···················· |
| A | 3194 hCoV-19/Australia/VIC718/2020\|EPI_ISL_427002\|2020- | ···················· | ···················· | ···················· | ···················· | ···················· | ···················· |
| A | 3198 hCoV-19/Australia/VIC731/2020\|EPI_ISL_427010\|2020- | ···················· | ···················· | ···················· | ···················· | ···················· | ···················· |
| A | 3201 hCoV-19/Australia/VIC734/2020\|EPI_ISL_427013\|2020- | ···················· | ···················· | ···················· | ···················· | ···················· | ···················· |
| A | 3207 hCoV-19/Australia/VIC741/2020\|EPI_ISL_427019\|2020- | ···················· | ···················· | ···················· | ···················· | ···················· | ···················· |
| A | 3215 hCoV-19/Australia/VIC752/2020\|EPI_ISL_427029\|2020- | ···················· | ···················· | ···················· | ···················· | ···················· | ···················· |
| A | 3221 hCoV-19/Greece/13/2020\|EPI_ISL_427043\|2020-03-18 | ···················· | ···················· | ···················· | ···················· | ···················· | ···················· |
| A | 3226 hCoV-19/USA/WI-UW-204/2020\|EPI_ISL_428300\|2020-03- | ···················· | ···················· | ···················· | ···················· | ···················· | ···················· |
| A | 3227 hCoV-19/USA/WI-UW-205/2020\|EPI_ISL_428301\|2020-03- | ···················· | ···················· | ···················· | ···················· | ···················· | ···················· |
| A | 3228 hCoV-19/USA/WI-UW-206/2020\|EPI_ISL_428302\|2020-03- | ···················· | ···················· | ···················· | ···················· | ···················· | ···················· |
| A | 3229 hCoV-19/USA/WI-UW-207/2020\|EPI_ISL_428303\|2020-03- | ···················· | ···················· | ···················· | ···················· | ···················· | ···················· |
| A | 3230 hCoV-19/USA/WI-UW-208/2020\|EPI_ISL_428304\|2020-03- | ···················· | ···················· | ···················· | ···················· | ···················· | ···················· |
| A | 3231 hCoV-19/USA/WI-UW-209/2020\|EPI_ISL_428305\|2020-03- | ···················· | ···················· | ···················· | ···················· | ···················· | ···················· |
| A | 3232 hCoV-19/USA/WI-UW-210/2020\|EPI_ISL_428306\|2020-03- | ···················· | ···················· | ···················· | ···················· | ···················· | ···················· |
| A | 3233 hCoV-19/USA/WI-UW-211/2020\|EPI_ISL_428307\|2020-03- | ···················· | ···················· | ···················· | ···················· | ···················· | ···················· |
| A | 3234 hCoV-19/USA/WI-UW-212/2020\|EPI_ISL_428308\|2020-03- | ···················· | ···················· | ···················· | ···················· | ···················· | ···················· |
| A | 3235 hCoV-19/USA/WI-UW-213/2020\|EPI_ISL_428309\|2020-03- | ···················· | ···················· | ···················· | ···················· | ···················· | ···················· |
| A | 3236 hCoV-19/USA/WI-UW-214/2020\|EPI_ISL_428310\|2020-03- | ···················· | ···················· | ···················· | ···················· | ···················· | ···················· |
| A | 3237 hCoV-19/USA/WI-UW-215/2020\|EPI_ISL_428311\|2020-03- | ···················· | ···················· | ···················· | ···················· | ···················· | ···················· |
| A | 3238 hCoV-19/USA/WI-UW-216/2020\|EPI_ISL_428312\|2020-03- | ···················· | ···················· | ···················· | ···················· | ···················· | ···················· |
| A | 3239 hCoV-19/USA/WI-UW-217/2020\|EPI_ISL_428313\|2020-03- | ···················· | ···················· | ···················· | ···················· | ···················· | ···················· |
| A | 3240 hCoV-19/USA/WI-UW-218/2020\|EPI_ISL_428314\|2020-03- | ···················· | ···················· | ···················· | ···················· | ···················· | ···················· |
| A | 3241 hCoV-19/USA/WI-UW-219/2020\|EPI_ISL_428315\|2020-03- | ···················· | ···················· | ···················· | ···················· | ···················· | ···················· |
| A | 3242 hCoV-19/USA/WI-UW-220/2020\|EPI_ISL_428316\|2020-03- | ···················· | ···················· | ···················· | ···················· | ···················· | ···················· |
| A | 3243 hCoV-19/USA/WI-UW-221/2020\|EPI_ISL_428317\|2020-03- | ···················· | ···················· | ···················· | ···················· | ···················· | ···················· |

| | | | | | | | | |
|---|---|---|---|---|---|---|---|---|
| A | 3244 hCoV-19/USA/WI-UW-222/2020\|EPI_ISL_428318\|2020-03- | .................... | .................... | .................... | .................... | .................... | .................... | |
| A | 3245 hCoV-19/USA/WI-UW-223/2020\|EPI_ISL_428319\|2020-03- | .................... | .................... | .................... | .................... | .................... | .................... | |
| A | 3246 hCoV-19/USA/WI-UW-224/2020\|EPI_ISL_428320\|2020-03- | .................... | .................... | .................... | .................... | .................... | .................... | |
| A | 3247 hCoV-19/USA/WI-UW-225/2020\|EPI_ISL_428321\|2020-03- | .................... | .................... | .................... | .................... | .................... | .................... | |
| A | 3248 hCoV-19/USA/WI-UW-226/2020\|EPI_ISL_428322\|2020-03- | .................... | .................... | .................... | .................... | .................... | .................... | |
| A | 3249 hCoV-19/USA/WI-UW-227/2020\|EPI_ISL_428323\|2020-03- | .................... | .................... | .................... | .................... | .................... | .................... | |
| A | 3250 hCoV-19/USA/WI-UW-228/2020\|EPI_ISL_428324\|2020-03- | .................... | .................... | .................... | .................... | .................... | .................... | |
| A | 3251 hCoV-19/USA/WI-UW-229/2020\|EPI_ISL_428325\|2020-03- | .................... | .................... | .................... | .................... | .................... | .................... | |
| A | 3252 hCoV-19/USA/WI-UW-230/2020\|EPI_ISL_428326\|2020-03- | .................... | .................... | .................... | .................... | .................... | .................... | |
| A | 3253 hCoV-19/USA/WI-UW-231/2020\|EPI_ISL_428327\|2020-03- | .................... | .................... | .................... | .................... | .................... | .................... | |
| A | 3254 hCoV-19/USA/WI-UW-232/2020\|EPI_ISL_428328\|2020-03- | .................... | .................... | .................... | .................... | .................... | .................... | |
| A | 3255 hCoV-19/USA/WI-UW-233/2020\|EPI_ISL_428329\|2020-03- | .................... | .................... | .................... | .................... | .................... | .................... | |
| A | 3256 hCoV-19/USA/WI-UW-234/2020\|EPI_ISL_428330\|2020-03- | .................... | .................... | .................... | .................... | .................... | .................... | |
| A | 3257 hCoV-19/USA/WI-UW-235/2020\|EPI_ISL_428331\|2020-03- | .................... | .................... | .................... | .................... | .................... | .................... | |
| A | 3258 hCoV-19/USA/WI-UW-236/2020\|EPI_ISL_428332\|2020-03- | .................... | .................... | .................... | .................... | .................... | .................... | |
| A | 3259 hCoV-19/USA/WI-UW-237/2020\|EPI_ISL_428333\|2020-03- | .................... | .................... | .................... | .................... | .................... | .................... | |
| A | 3260 hCoV-19/USA/WI-UW-238/2020\|EPI_ISL_428334\|2020-03- | .................... | .................... | .................... | .................... | .................... | .................... | |
| A | 3261 hCoV-19/USA/WI-UW-239/2020\|EPI_ISL_428335\|2020-03- | .................... | .................... | .................... | .................... | .................... | .................... | |
| A | 3262 hCoV-19/USA/WI-UW-240/2020\|EPI_ISL_428336\|2020-03- | .................... | .................... | .................... | .................... | .................... | .................... | |
| A | 3263 hCoV-19/USA/WI-UW-241/2020\|EPI_ISL_428337\|2020-03- | .................... | .................... | .................... | .................... | .................... | .................... | |
| A | 3264 hCoV-19/USA/WI-UW-242/2020\|EPI_ISL_428338\|2020-03- | .................... | .................... | .................... | .................... | .................... | .................... | |
| A | 3265 hCoV-19/USA/WI-UW-243/2020\|EPI_ISL_428339\|2020-03- | .................... | .................... | .................... | .................... | .................... | .................... | |
| A | 3266 hCoV-19/USA/WI-UW-244/2020\|EPI_ISL_428340\|2020-03- | .................... | .................... | .................... | .................... | .................... | .................... | |
| A | 3267 hCoV-19/USA/WI-UW-245/2020\|EPI_ISL_428341\|2020-03- | .................... | .................... | .................... | .................... | .................... | .................... | |
| A | 3268 hCoV-19/USA/WI-UW-246/2020\|EPI_ISL_428342\|2020-03- | .................... | .................... | .................... | .................... | .................... | .................... | |
| A | 3269 hCoV-19/USA/WI-UW-247/2020\|EPI_ISL_428343\|2020-03- | .................... | .................... | .................... | .................... | .................... | .................... | |
| A | 3270 hCoV-19/USA/WI-UW-248/2020\|EPI_ISL_428344\|2020-03- | .................... | .................... | .................... | .................... | .................... | .................... | |
| A | 3271 hCoV-19/USA/WI-UW-249/2020\|EPI_ISL_428345\|2020-03- | .................... | .................... | .................... | .................... | .................... | .................... | |
| A | 3272 hCoV-19/Turkey/IST-GLAB-012/2020\|EPI_ISL_428346\|20 | .................... | .................... | .................... | .................... | .................... | .................... | |
| A | 3273 hCoV-19/France/IDF-3384/2020\|EPI_ISL_428347\|2020-0 | .................... | .................... | .................... | .................... | .................... | .................... | |
| A | 3274 hCoV-19/France/GES-3372/2020\|EPI_ISL_428348\|2020-0 | .................... | .................... | .................... | .................... | .................... | .................... | |
| A | 3275 hCoV-19/France/IDF-3386/2020\|EPI_ISL_428349\|2020-0 | .................... | .................... | .................... | .................... | .................... | .................... | |
| A | 3276 hCoV-19/France/HDF-3419/2020\|EPI_ISL_428350\|2020-0 | .................... | .................... | .................... | .................... | .................... | .................... | |
| A | 3277 hCoV-19/France/IDF-3509/2020\|EPI_ISL_428351\|2020-0 | .................... | .................... | .................... | .................... | .................... | .................... | |
| A | 3278 hCoV-19/France/IDF-3518/2020\|EPI_ISL_428352\|2020-0 | .................... | .................... | .................... | .................... | .................... | .................... | |
| A | 3279 hCoV-19/France/HDF-3534/2020\|EPI_ISL_428353\|2020-0 | .................... | .................... | .................... | .................... | .................... | .................... | |
| A | 3280 hCoV-19/France/IDF-3577/2020\|EPI_ISL_428354\|2020-0 | .................... | .................... | .................... | .................... | .................... | .................... | |
| A | 3281 hCoV-19/France/IDF-2251/2020\|EPI_ISL_428355\|2020-0 | .................... | .................... | .................... | .................... | .................... | .................... | |
| A | 3282 hCoV-19/France/IDF-2252/2020\|EPI_ISL_428356\|2020-0 | .................... | .................... | .................... | .................... | .................... | .................... | |
| A | 3283 hCoV-19/France/IDF-2253/2020\|EPI_ISL_428357\|2020-0 | .................... | .................... | .................... | .................... | .................... | .................... | |
| A | 3284 hCoV-19/France/HDF-3598/2020\|EPI_ISL_428358\|2020-0 | .................... | .................... | .................... | .................... | .................... | .................... | |
| A | 3285 hCoV-19/France/HDF-3677/2020\|EPI_ISL_428359\|2020-0 | .................... | .................... | .................... | .................... | .................... | .................... | |
| A | 3286 hCoV-19/France/HDF-3678/2020\|EPI_ISL_428360\|2020-0 | .................... | .................... | .................... | .................... | .................... | .................... | |
| A | 3287 hCoV-19/France/IDF-3703/2020\|EPI_ISL_428361\|2020-0 | .................... | .................... | .................... | .................... | .................... | .................... | |
| A | 3288 hCoV-19/France/IDF-3745/2020\|EPI_ISL_428363\|2020-0 | .................... | .................... | .................... | .................... | .................... | .................... | |
| A | 3289 hCoV-19/France/IDF-3930/2020\|EPI_ISL_428365\|2020-0 | .................... | .................... | .................... | .................... | .................... | .................... | |
| A | 3290 hCoV-19/France/HDF-4220/2020\|EPI_ISL_428366\|2020-0 | .................... | .................... | .................... | .................... | .................... | .................... | |
| A | 3291 hCoV-19/France/NOR-N4427/2020\|EPI_ISL_428367\|2020- | .................... | .................... | .................... | .................... | .................... | .................... | |
| A | 3292 hCoV-19/Turkey/IST-GLAB-013/2020\|EPI_ISL_428368\|20 | .................... | .................... | .................... | .................... | .................... | .................... | |
| A | 3293 hCoV-19/USA/WA-UW-4649/2020\|EPI_ISL_429629\|2020-04 | .................... | .................... | .................... | .................... | .................... | .................... | |
| A | 3294 hCoV-19/USA/WA-UW-4677/2020\|EPI_ISL_429630\|2020-04 | .................... | .................... | .................... | .................... | .................... | .................... | |
| A | 3295 hCoV-19/USA/WA-UW-4695/2020\|EPI_ISL_429631\|2020-04 | .................... | .................... | .................... | .................... | .................... | .................... | |
| A | 3296 hCoV-19/USA/WA-UW-4705/2020\|EPI_ISL_429632\|2020-04 | .................... | .................... | .................... | .................... | .................... | .................... | |
| A | 3297 hCoV-19/USA/WA-UW-4892/2020\|EPI_ISL_429634\|2020-04 | .................... | .................... | .................... | .................... | .................... | .................... | |
| A | 3298 hCoV-19/USA/WA-UW-4897/2020\|EPI_ISL_429635\|2020-04 | .................... | .................... | .................... | .................... | .................... | .................... | |
| A | 3299 hCoV-19/USA/WA-UW-4921/2020\|EPI_ISL_429636\|2020-04 | .................... | .................... | .................... | .................... | .................... | .................... | |
| A | 3300 hCoV-19/USA/WA-UW-4925/2020\|EPI_ISL_429637\|2020-04 | .................... | .................... | .................... | .................... | .................... | .................... | |
| A | 3301 hCoV-19/USA/WA-UW-4999/2020\|EPI_ISL_429638\|2020-04 | .................... | .................... | .................... | .................... | .................... | .................... | |
| A | 3302 hCoV-19/USA/WA-UW-5025/2020\|EPI_ISL_429639\|2020-04 | .................... | .................... | .................... | .................... | .................... | .................... | |
| A | 3303 hCoV-19/USA/CT-UW-5038/2020\|EPI_ISL_429641\|2020-04 | .................... | .................... | .................... | .................... | .................... | .................... | |
| A | 3304 hCoV-19/USA/CT-UW-5043/2020\|EPI_ISL_429642\|2020-04 | .................... | ...................A | .................... | .................... | .................... | .................... | 1 |
| A | 3305 hCoV-19/USA/UN-UW-5044/2020\|EPI_ISL_429643\|2020-04 | .................... | .................... | .................... | .................... | .................... | .................... | |
| A | 3306 hCoV-19/USA/WA-UW-5065/2020\|EPI_ISL_429644\|2020-04 | .................... | .................... | .................... | .................... | .................... | .................... | |
| A | 3307 hCoV-19/USA/CT-UW-5073/2020\|EPI_ISL_429645\|2020-04 | .................... | .................... | .................... | .................... | .................... | .................... | |
| A | 3308 hCoV-19/USA/WA-UW-5095/2020\|EPI_ISL_429647\|2020-04 | .................... | .................... | .................... | .................... | .................... | .................... | |
| A | 3309 hCoV-19/USA/WA-UW-5144/2020\|EPI_ISL_429649\|2020-04 | .................... | .................... | .................... | .................... | .................... | .................... | |
| A | 3310 hCoV-19/USA/WA-UW-5152/2020\|EPI_ISL_429650\|2020-04 | .................... | .................... | .................... | .................... | .................... | .................... | |
| A | 3311 hCoV-19/USA/WA-UW-5158/2020\|EPI_ISL_429651\|2020-04 | .................... | .................... | .................... | .................... | .................... | .................... | |
| A | 3312 hCoV-19/USA/WA-UW-5164/2020\|EPI_ISL_429652\|2020-04 | .................... | .................... | .................... | .................... | .................... | .................... | |
| A | 3313 hCoV-19/USA/WA-UW-5166/2020\|EPI_ISL_429653\|2020-04 | .................... | .................... | .................... | .................... | .................... | .................... | |
| A | 3314 hCoV-19/USA/WA-UW-5169/2020\|EPI_ISL_429654\|2020-04 | .................... | .................... | .................... | .................... | .................... | .................... | |
| A | 3315 hCoV-19/USA/WA-UW-5170/2020\|EPI_ISL_429655\|2020-04 | .................... | .................... | .................... | .................... | .................... | .................... | |
| A | 3316 hCoV-19/USA/UN-UW-5172/2020\|EPI_ISL_429656\|2020-04 | .................... | .................... | .................... | .................... | .................... | .................... | |
| A | 3317 hCoV-19/Croatia/OY-S1/2020\|EPI_ISL_429659\|2020-03 | .................... | .................... | .................... | .................... | .................... | .................... | |
| A | 3318 hCoV-19/Croatia/8U-S17/2020\|EPI_ISL_429663\|2020-03 | .................... | .................... | .................... | .................... | .................... | .................... | |
| A | 3319 hCoV-19/Brazil/MG-CV4/2020\|EPI_ISL_429667\|2020-03- | .................... | .................... | .................... | .................... | .................... | .................... | |
| A | 3320 hCoV-19/Brazil/MG-CV6/2020\|EPI_ISL_429669\|2020-03- | .................... | .................... | .................... | .................... | .................... | .................... | |
| A | 3321 hCoV-19/Brazil/MG-CV8/2020\|EPI_ISL_429671\|2020-03- | .................... | .................... | .................... | .................... | .................... | .................... | |

| | | | | | | | | |
|---|---|---|---|---|---|---|---|---|
| A | 3322 | hCoV-19/Brazil/MG-CV12/2020\|EPI_ISL_429674\|2020-03 | .................... | .................... | ..................... | .................... | .................... | .................... |
| A | 3323 | hCoV-19/Brazil/MG-CV16/2020\|EPI_ISL_429676\|2020-03 | .................... | .................... | ..................... | .................... | .................... | .................... |
| A | 3324 | hCoV-19/Brazil/MG-CV21/2020\|EPI_ISL_429681\|2020-03 | .................... | .................... | ..................... | .................... | .................... | .................... |
| A | 3325 | hCoV-19/Brazil/MG-CV31/2020\|EPI_ISL_429687\|2020-03 | .................... | .................... | ..................... | .................... | .................... | .................... |
| A | 3326 | hCoV-19/Brazil/MG-CV32/2020\|EPI_ISL_429688\|2020-03 | .................... | .................... | ..................... | .................... | .................... | .................... |
| A | 3327 | hCoV-19/Brazil/MG-CV33/2020\|EPI_ISL_429689\|2020-03 | .................... | .................... | ..................... | .................... | .................... | .................... |
| A | 3328 | hCoV-19/Brazil/MG-CV42/2020\|EPI_ISL_429695\|2020-03 | .................... | .................... | ..................... | .................... | .................... | .................... |
| A | 3329 | hCoV-19/Hangzhou/HZ-1/2020\|EPI_ISL_406970\|2020-01- | .................... | .................... | ..................... | .................... | .................... | .................... |
| A | 3330 | hCoV-19/Singapore/1/2020\|EPI_ISL_406973\|2020-01-23 | .................... | .................... | ..................... | .................... | .................... | .................... |
| A | 3331 | hCoV-19/Hong Kong/CUHK1/2020\|EPI_ISL_416314\|2020-0 | .................... | .................... | ..................... | .................... | .................... | .................... |
| A | 3332 | hCoV-19/Shanghai/SH0002/2020\|EPI_ISL_416316\|2020-0 | .................... | .................... | ..................... | .................... | .................... | .................... |
| A | 3333 | hCoV-19/Shanghai/SH0003/2020\|EPI_ISL_416317\|2020-0 | .................... | .................... | ..................... | .................... | .................... | .................... |
| A | 3334 | hCoV-19/Shanghai/SH0004/2020\|EPI_ISL_416318\|2020-0 | .................... | .................... | ..................... | .................... | .................... | .................... |
| A | 3335 | hCoV-19/Iceland/272/2020\|EPI_ISL_417600\|2020-03-17 | .................... | .................... | ..................... | .................... | .................... | .................... |
| A | 3336 | hCoV-19/Iceland/273/2020\|EPI_ISL_417601\|2020-03-17 | .................... | .................... | ..................... | .................... | .................... | .................... |
| A | 3337 | hCoV-19/Iceland/274/2020\|EPI_ISL_417602\|2020-03-17 | .................... | .................... | ..................... | .................... | .................... | .................... |
| A | 3338 | hCoV-19/Iceland/275/2020\|EPI_ISL_417603\|2020-03-17 | .................... | .................... | ..................... | .................... | .................... | .................... |
| A | 3339 | hCoV-19/Iceland/276/2020\|EPI_ISL_417604\|2020-03-17 | .................... | .................... | ..................... | .................... | .................... | .................... |
| A | 3340 | hCoV-19/Iceland/277/2020\|EPI_ISL_417605\|2020-03-17 | .................... | .................... | ..................... | .................... | .................... | .................... |
| A | 3341 | hCoV-19/Iceland/278/2020\|EPI_ISL_417606\|2020-03-17 | .................... | .................... | ..................... | .................... | .................... | .................... |
| A | 3342 | hCoV-19/Iceland/279/2020\|EPI_ISL_417607\|2020-03-17 | .................... | .................... | ..................... | .................... | .................... | .................... |
| A | 3343 | hCoV-19/Iceland/280/2020\|EPI_ISL_417608\|2020-03-17 | .................... | .................... | ..................... | .................... | .................... | .................... |
| A | 3344 | hCoV-19/Iceland/281/2020\|EPI_ISL_417609\|2020-03-17 | .................... | .................... | ..................... | .................... | .................... | .................... |
| A | 3345 | hCoV-19/Iceland/282/2020\|EPI_ISL_417610\|2020-03-17 | .................... | .................... | ..................... | .................... | .................... | .................... |
| A | 3346 | hCoV-19/Iceland/283/2020\|EPI_ISL_417611\|2020-03-17 | .................... | .................... | ..................... | .................... | .................... | .................... |
| A | 3347 | hCoV-19/Iceland/284/2020\|EPI_ISL_417612\|2020-03-17 | .................... | .................... | ..................... | .................... | .................... | .................... |
| A | 3348 | hCoV-19/Iceland/285/2020\|EPI_ISL_417613\|2020-03-17 | .................... | .................... | ..................... | .................... | .................... | .................... |
| A | 3349 | hCoV-19/Iceland/286/2020\|EPI_ISL_417614\|2020-03-18 | .................... | .................... | ..................... | .................... | .................... | .................... |
| A | 3350 | hCoV-19/Iceland/287/2020\|EPI_ISL_417615\|2020-03-18 | .................... | .................... | ..................... | .................... | .................... | .................... |
| A | 3351 | hCoV-19/Iceland/288/2020\|EPI_ISL_417616\|2020-03-18 | .................... | .................... | ..................... | .................... | .................... | .................... |
| A | 3352 | hCoV-19/Iceland/289/2020\|EPI_ISL_417617\|2020-03-18 | .................... | .................... | ..................... | .................... | .................... | .................... |
| A | 3353 | hCoV-19/Iceland/29/2020\|EPI_ISL_417618\|2020-03-16 | .................... | .................... | ..................... | .................... | .................... | .................... |
| A | 3354 | hCoV-19/Iceland/290/2020\|EPI_ISL_417619\|2020-03-18 | .................... | .................... | ..................... | .................... | .................... | .................... |
| A | 3355 | hCoV-19/Iceland/291/2020\|EPI_ISL_417620\|2020-03-18 | .................... | .................... | ..................... | .................... | .................... | .................... |
| A | 3356 | hCoV-19/Iceland/292/2020\|EPI_ISL_417621\|2020-03-18 | .................... | .................... | ..................... | .................... | .................... | .................... |
| A | 3357 | hCoV-19/Iceland/293/2020\|EPI_ISL_417622\|2020-03-18 | .................... | .................... | ..................... | .................... | .................... | .................... |
| A | 3358 | hCoV-19/Iceland/294/2020\|EPI_ISL_417623\|2020-03-18 | .................... | .................... | ..................... | .................... | .................... | .................... |
| A | 3359 | hCoV-19/Iceland/295/2020\|EPI_ISL_417624\|2020-03-18 | .................... | .................... | ..................... | .................... | .................... | .................... |
| A | 3360 | hCoV-19/Iceland/296/2020\|EPI_ISL_417625\|2020-03-18 | .................... | .................... | ..................... | .................... | .................... | .................... |
| A | 3361 | hCoV-19/Iceland/297/2020\|EPI_ISL_417626\|2020-03-18 | .................... | .................... | ..................... | .................... | .................... | .................... |
| A | 3362 | hCoV-19/Iceland/298/2020\|EPI_ISL_417627\|2020-03-18 | .................... | .................... | ..................... | .................... | .................... | .................... |
| A | 3363 | hCoV-19/Iceland/299/2020\|EPI_ISL_417628\|2020-03-18 | .................... | .................... | ..................... | .................... | .................... | .................... |
| A | 3364 | hCoV-19/Iceland/300/2020\|EPI_ISL_417629\|2020-03-18 | .................... | .................... | ..................... | .................... | .................... | .................... |
| A | 3365 | hCoV-19/Iceland/301/2020\|EPI_ISL_417630\|2020-03-18 | .................... | .................... | ..................... | .................... | .................... | .................... |
| A | 3366 | hCoV-19/Iceland/302/2020\|EPI_ISL_417631\|2020-03-18 | .................... | .................... | ..................... | .................... | .................... | .................... |
| A | 3367 | hCoV-19/Iceland/303/2020\|EPI_ISL_417632\|2020-03-18 | .................... | .................... | ..................... | .................... | .................... | .................... |
| A | 3368 | hCoV-19/Iceland/304/2020\|EPI_ISL_417633\|2020-03-17 | .................... | .................... | ..................... | .................... | .................... | .................... |
| A | 3369 | hCoV-19/Iceland/305/2020\|EPI_ISL_417634\|2020-03-18 | .................... | .................... | ..................... | .................... | .................... | .................... |
| A | 3370 | hCoV-19/Iceland/306/2020\|EPI_ISL_417635\|2020-03-18 | .................... | .................... | ..................... | .................... | .................... | .................... |
| A | 3371 | hCoV-19/Iceland/307/2020\|EPI_ISL_417636\|2020-03-18 | .................... | .................... | ..................... | .................... | .................... | .................... |
| A | 3372 | hCoV-19/Iceland/308/2020\|EPI_ISL_417637\|2020-03-18 | .................... | .................... | ..................... | .................... | .................... | .................... |
| A | 3373 | hCoV-19/Iceland/309/2020\|EPI_ISL_417638\|2020-03-18 | .................... | .................... | ..................... | .................... | .................... | .................... |
| A | 3374 | hCoV-19/Iceland/310/2020\|EPI_ISL_417639\|2020-03-18 | .................... | .................... | ..................... | .................... | .................... | .................... |
| A | 3375 | hCoV-19/Iceland/311/2020\|EPI_ISL_417640\|2020-03-18 | .................... | .................... | ..................... | .................... | .................... | .................... |
| A | 3376 | hCoV-19/Iceland/312/2020\|EPI_ISL_417641\|2020-03-18 | .................... | .................... | ..................... | .................... | .................... | .................... |
| A | 3377 | hCoV-19/Iceland/313/2020\|EPI_ISL_417642\|2020-03-18 | .................... | .................... | ..................... | .................... | .................... | .................... |
| A | 3378 | hCoV-19/Iceland/314/2020\|EPI_ISL_417643\|2020-03-18 | .................... | .................... | ..................... | .................... | .................... | .................... |
| A | 3379 | hCoV-19/Iceland/315/2020\|EPI_ISL_417644\|2020-03-18 | .................... | .................... | ..................... | .................... | .................... | .................... |
| A | 3380 | hCoV-19/Iceland/316/2020\|EPI_ISL_417645\|2020-03-18 | .................... | .................... | ..................... | .................... | .................... | .................... |
| A | 3381 | hCoV-19/Iceland/317/2020\|EPI_ISL_417646\|2020-03-18 | .................... | .................... | ..................... | .................... | .................... | .................... |
| A | 3382 | hCoV-19/Iceland/318/2020\|EPI_ISL_417647\|2020-03-18 | .................... | .................... | ..................... | .................... | .................... | .................... |
| A | 3383 | hCoV-19/Iceland/319/2020\|EPI_ISL_417648\|2020-03-18 | .................... | .................... | ..................... | .................... | .................... | .................... |
| A | 3384 | hCoV-19/Iceland/320/2020\|EPI_ISL_417649\|2020-03-18 | .................... | .................... | ..................... | .................... | .................... | .................... |
| A | 3385 | hCoV-19/USA/WA-UW333/2020\|EPI_ISL_418900\|2020-03-1 | .................... | .................... | ..................... | .................... | .................... | .................... |
| A | 3386 | hCoV-19/USA/MN-UW334/2020\|EPI_ISL_418901\|2020-03-1 | .................... | .................... | ..................... | .................... | .................... | .................... |
| A | 3387 | hCoV-19/USA/MN-UW335/2020\|EPI_ISL_418902\|2020-03-1 | .................... | .................... | ..................... | .................... | .................... | .................... |
| A | 3388 | hCoV-19/USA/WA-UW336/2020\|EPI_ISL_418903\|2020-03-1 | .................... | .................... | ..................... | .................... | .................... | .................... |
| A | 3389 | hCoV-19/USA/WA-UW337/2020\|EPI_ISL_418904\|2020-03-1 | .................... | .................... | ..................... | .................... | .................... | .................... |
| A | 3390 | hCoV-19/USA/WA-UW338/2020\|EPI_ISL_418905\|2020-03-1 | .................... | .................... | ..................... | .................... | .................... | .................... |
| A | 3391 | hCoV-19/USA/WA-UW339/2020\|EPI_ISL_418906\|2020-03-1 | .................... | .................... | ..................... | .................... | .................... | .................... |
| A | 3392 | hCoV-19/USA/WA-UW340/2020\|EPI_ISL_418907\|2020-03-1 | .................... | .................... | ..................... | .................... | .................... | .................... |
| A | 3393 | hCoV-19/USA/WA-UW341/2020\|EPI_ISL_418908\|2020-03-1 | .................... | .................... | ..................... | .................... | .................... | .................... |
| A | 3394 | hCoV-19/USA/WA-UW342/2020\|EPI_ISL_418909\|2020-03-1 | .................... | .................... | ..................... | .................... | .................... | .................... |
| A | 3395 | hCoV-19/USA/WA-UW343/2020\|EPI_ISL_418910\|2020-03-1 | .................... | .................... | ..................... | .................... | .................... | .................... |
| A | 3396 | hCoV-19/USA/WA-UW344/2020\|EPI_ISL_418911\|2020-03-1 | .................... | .................... | ..................... | .................... | .................... | .................... |
| A | 3397 | hCoV-19/USA/WA-UW345/2020\|EPI_ISL_418912\|2020-03-1 | .................... | .................... | ..................... | .................... | .................... | .................... |
| A | 3398 | hCoV-19/USA/WA-UW346/2020\|EPI_ISL_418913\|2020-03-1 | .................... | .................... | ..................... | .................... | .................... | .................... |
| A | 3399 | hCoV-19/USA/IL-UW348/2020\|EPI_ISL_418915\|2020-03-1 | .................... | .................... | ..................... | .................... | .................... | .................... |

| | | | | | | | | |
|---|---|---|---|---|---|---|---|---|
| A | 3400 | hCoV-19/USA/IL-UW349/2020\|EPI_ISL_418916\|2020-03-1 | ···················· | ·A·················· | ···················· | ···················· | ···················· | ···················· |
| A | 3401 | hCoV-19/USA/IL-UW350/2020\|EPI_ISL_418917\|2020-03-1 | ···················· | ···················· | ···················· | ···················· | ···················· | ···················· |
| A | 3402 | hCoV-19/USA/IL-UW351/2020\|EPI_ISL_418918\|2020-03-1 | ···················· | ···················· | ···················· | ···················· | ···················· | ···················· |
| A | 3403 | hCoV-19/USA/IL-UW352/2020\|EPI_ISL_418919\|2020-03-1 | ···················· | ···················· | ···················· | ···················· | ···················· | ···················· |
| A | 3404 | hCoV-19/USA/IL-UW353/2020\|EPI_ISL_418920\|2020-03-1 | ···················· | ···················· | ···················· | ···················· | ···················· | ···················· |
| A | 3405 | hCoV-19/USA/UN-UW354/2020\|EPI_ISL_418921\|2020-03-1 | ···················· | ···················· | ···················· | ···················· | ···················· | ···················· |
| A | 3406 | hCoV-19/USA/WA-UW355/2020\|EPI_ISL_418922\|2020-03-1 | ···················· | ···················· | ···················· | ···················· | ···················· | ···················· |
| A | 3407 | hCoV-19/USA/WA-UW356/2020\|EPI_ISL_418923\|2020-03-1 | ···················· | ···················· | ···················· | ···················· | ···················· | ···················· |
| A | 3408 | hCoV-19/USA/WA-UW357/2020\|EPI_ISL_418924\|2020-03-1 | ···················· | ···················· | ···················· | ···················· | ···················· | ···················· |
| A | 3409 | hCoV-19/USA/UN-UW358/2020\|EPI_ISL_418925\|2020-03 | ···················· | ···················· | ···················· | ···················· | ···················· | ···················· |
| A | 3410 | hCoV-19/USA/WA-UW359/2020\|EPI_ISL_418926\|2020-03-1 | ···················· | ···················· | ···················· | ···················· | ···················· | ···················· |
| A | 3411 | hCoV-19/USA/WA-UW360/2020\|EPI_ISL_418927\|2020-03-1 | ···················· | ···················· | ···················· | ···················· | ···················· | ···················· |
| A | 3412 | hCoV-19/USA/WA-UW361/2020\|EPI_ISL_418928\|2020-03-1 | ···················· | ···················· | ···················· | ···················· | ···················· | ···················· |
| A | 3413 | hCoV-19/USA/WA-UW362/2020\|EPI_ISL_418929\|2020-03-1 | ···················· | ···················· | ···················· | ···················· | ···················· | ···················· |
| A | 3414 | hCoV-19/USA/WA-UW363/2020\|EPI_ISL_418930\|2020-03-1 | ···················· | ···················· | ···················· | ···················· | ···················· | ···················· |
| A | 3415 | hCoV-19/USA/WA-UW364/2020\|EPI_ISL_418931\|2020-03-1 | ···················· | ···················· | ···················· | ···················· | ···················· | ···················· |
| A | 3416 | hCoV-19/USA/WA-UW365/2020\|EPI_ISL_418932\|2020-03-1 | ···················· | ···················· | ···················· | ···················· | ···················· | ···················· |
| A | 3417 | hCoV-19/USA/WA-UW366/2020\|EPI_ISL_418933\|2020-03-1 | ···················· | ···················· | ···················· | ···················· | ···················· | ···················· |
| A | 3418 | hCoV-19/USA/WA-UW367/2020\|EPI_ISL_418934\|2020-03-1 | ···················· | ···················· | ···················· | ···················· | ···················· | ···················· |
| A | 3419 | hCoV-19/USA/WA-UW368/2020\|EPI_ISL_418935\|2020-03-1 | ···················· | ···················· | ···················· | ···················· | ···················· | ···················· |
| A | 3420 | hCoV-19/USA/WA-UW369/2020\|EPI_ISL_418936\|2020-03-1 | ···················· | ···················· | ···················· | ···················· | ···················· | ···················· |
| A | 3421 | hCoV-19/USA/WA-UW370/2020\|EPI_ISL_418937\|2020-03-1 | ···················· | ···················· | ···················· | ···················· | ···················· | ···················· |
| A | 3422 | hCoV-19/USA/WA-UW371/2020\|EPI_ISL_418938\|2020-03-1 | ···················· | ···················· | ···················· | ···················· | ···················· | ···················· |
| A | 3423 | hCoV-19/USA/WA-UW372/2020\|EPI_ISL_418939\|2020-03-1 | ···················· | ···················· | ···················· | ···················· | ···················· | ···················· |
| A | 3424 | hCoV-19/USA/WA-UW373/2020\|EPI_ISL_418940\|2020-03-1 | ···················· | ···················· | ···················· | ···················· | ···················· | ···················· |
| A | 3425 | hCoV-19/USA/WA-UW374/2020\|EPI_ISL_418941\|2020-03 | ···················· | ···················· | ···················· | ···················· | ···················· | ···················· |
| A | 3426 | hCoV-19/USA/WA-UW375/2020\|EPI_ISL_418942\|2020-03-1 | ···················· | ···················· | ···················· | ···················· | ···················· | ···················· |
| A | 3427 | hCoV-19/USA/WA-UW376/2020\|EPI_ISL_418943\|2020-03-1 | ···················· | ···················· | ···················· | ···················· | ···················· | ···················· |
| A | 3428 | hCoV-19/USA/WA-UW378/2020\|EPI_ISL_418944\|2020-03-1 | ···················· | ···················· | ···················· | ···················· | ···················· | ···················· |
| A | 3429 | hCoV-19/USA/WA-UW-1238/2020\|EPI_ISL_418945\|2020-03 | ···················· | ···················· | ···················· | ···················· | ···················· | ···················· |
| A | 3430 | hCoV-19/USA/WA-UW383/2020\|EPI_ISL_418948\|2020-03-1 | ···················· | ···················· | ···················· | ···················· | ···················· | ···················· |
| A | 3431 | hCoV-19/USA/WA-UW384/2020\|EPI_ISL_418949\|2020-03-1 | ···················· | ···················· | ···················· | ···················· | ···················· | ···················· |
| A | 3432 | hCoV-19/USA/WA-UW385/2020\|EPI_ISL_418950\|2020-03-1 | ···················· | ···················· | ···················· | ···················· | ···················· | ···················· |
| A | 3433 | hCoV-19/USA/WA-UW386/2020\|EPI_ISL_418951\|2020-03-1 | ···················· | ···················· | ···················· | ···················· | ···················· | ···················· |
| A | 3434 | hCoV-19/USA/WA-UW387/2020\|EPI_ISL_418952\|2020-03-1 | ···················· | ···················· | ···················· | ···················· | ···················· | ···················· |
| A | 3435 | hCoV-19/USA/WA-UW388/2020\|EPI_ISL_418953\|2020-03-1 | ···················· | ···················· | ···················· | ···················· | ···················· | ···················· |
| A | 3436 | hCoV-19/USA/WA-UW389/2020\|EPI_ISL_418954\|2020-03-1 | ···················· | ···················· | ···················· | ···················· | ···················· | ···················· |
| A | 3437 | hCoV-19/USA/WA-UW390/2020\|EPI_ISL_418955\|2020-03-1 | ···················· | ···················· | ···················· | ···················· | ···················· | ···················· |
| A | 3438 | hCoV-19/USA/VA-DCLS-0002/2020\|EPI_ISL_418956\|2020- | ···················· | ···················· | ···················· | ···················· | ···················· | ···················· |
| A | 3439 | hCoV-19/USA/VA-DCLS-0003/2020\|EPI_ISL_418957\|2020- | ···················· | ···················· | ···················· | ···················· | ···················· | ···················· |
| A | 3440 | hCoV-19/USA/VA-DCLS-0004/2020\|EPI_ISL_418958\|2020- | ···················· | ···················· | ···················· | ···················· | ···················· | ···················· |
| A | 3441 | hCoV-19/USA/UT-019/2020\|EPI_ISL_418962\|2020-03-13 | ···················· | ···················· | ···················· | ···················· | ···················· | ···················· |
| A | 3442 | hCoV-19/USA/UT-021/2020\|EPI_ISL_418963\|2020-03-14 | ···················· | ···················· | ···················· | ···················· | ···················· | ···················· |
| A | 3443 | hCoV-19/USA/UT-022/2020\|EPI_ISL_418964\|2020-03-14 | ···················· | ···················· | ···················· | ···················· | ···················· | ···················· |
| A | 3444 | hCoV-19/USA/UT-023/2020\|EPI_ISL_418965\|2020-03-11 | ···················· | ···················· | ···················· | ···················· | ···················· | ···················· |
| A | 3445 | hCoV-19/USA/UT-025/2020\|EPI_ISL_418967\|2020-03-11 | ···················· | ···················· | ···················· | ···················· | ···················· | ···················· |
| A | 3446 | hCoV-19/USA/NY-NYUMC26/2020\|EPI_ISL_418972\|2020-03 | ···················· | ···················· | ···················· | ···················· | ···················· | ···················· |
| A | 3447 | hCoV-19/USA/NY-NYUMC27/2020\|EPI_ISL_418973\|2020-03 | ···················· | ···················· | ···················· | ···················· | ···················· | ···················· |
| A | 3448 | hCoV-19/USA/NY-NYUMC28/2020\|EPI_ISL_418974\|2020-03 | ···················· | ···················· | ···················· | ···················· | ···················· | ···················· |
| A | 3449 | hCoV-19/USA/NY-NYUMC29/2020\|EPI_ISL_418975\|2020-03 | ···················· | ···················· | ···················· | ···················· | ···················· | ···················· |
| A | 3450 | hCoV-19/USA/NY-NYUMC30/2020\|EPI_ISL_418976\|2020-03 | ···················· | ···················· | ···················· | ···················· | ···················· | ···················· |
| A | 3451 | hCoV-19/USA/NY-NYUMC31/2020\|EPI_ISL_418977\|2020-03 | ···················· | ···················· | ···················· | ···················· | ···················· | ···················· |
| A | 3452 | hCoV-19/USA/NY-NYUMC32/2020\|EPI_ISL_418978\|2020-03 | ···················· | ···················· | ···················· | ···················· | ···················· | ···················· |
| A | 3453 | hCoV-19/USA/NY-NYUMC33/2020\|EPI_ISL_418979\|2020-03 | ···················· | ···················· | ···················· | NN·················· | ···················· | ···················· |
| A | 3454 | hCoV-19/Wales/PHWC-260B9/2020\|EPI_ISL_422100\|2020- | ···················· | ···················· | ···················· | ···················· | ···················· | ···················· |
| A | 3455 | hCoV-19/Wales/PHWC-26444/2020\|EPI_ISL_422102\|2020- | ···················· | ···················· | ···················· | ···················· | ···················· | ···················· |
| A | 3456 | hCoV-19/Wales/PHWC-26C00/2020\|EPI_ISL_422103\|2020- | ········Y··········· | ···················· | ···················· | ···················· | ···················· | ···················· |
| A | 3457 | hCoV-19/Wales/PHWC-26022/2020\|EPI_ISL_422104\|2020- | ···················· | ····A··············· | ···················· | ···················· | ···················· | ···················· |
| A | 3458 | hCoV-19/Wales/PHWC-26F25/2020\|EPI_ISL_422105\|2020- | ···················· | ···················· | ···················· | ···················· | ···················· | ···················· |
| A | 3459 | hCoV-19/Wales/PHWC-260F5/2020\|EPI_ISL_422106\|2020- | ···················· | ····A··············· | ···················· | ···················· | ···················· | ···················· |
| A | 3460 | hCoV-19/Wales/PHWC-25FF9/2020\|EPI_ISL_422107\|2020- | ···················· | ···················· | ···················· | ···················· | ···················· | ···················· |
| A | 3461 | hCoV-19/Wales/PHWC-2581C/2020\|EPI_ISL_422108\|2020- | ···················· | ···················· | ···················· | ···················· | ···················· | ···················· |
| A | 3462 | hCoV-19/Wales/PHWC-266F3/2020\|EPI_ISL_422109\|2020- | ···················· | ···················· | ···················· | ···················· | ···················· | ···················· |
| A | 3463 | hCoV-19/Wales/PHWC-260D7/2020\|EPI_ISL_422110\|2020- | ···················· | ···················· | ···················· | ···················· | ···················· | ···················· |
| A | 3464 | hCoV-19/Wales/PHWC-260C8/2020\|EPI_ISL_422111\|2020- | ···················· | ···················· | ···················· | ···················· | ···················· | ···················· |
| A | 3465 | hCoV-19/Wales/PHWC-26417/2020\|EPI_ISL_422112\|2020- | ···················· | ···················· | ···················· | ···················· | ···················· | ···················· |
| A | 3466 | hCoV-19/Wales/PHWC-26CB5/2020\|EPI_ISL_422113\|2020- | ···················· | ····M··············· | ···················· | ···················· | ···················· | ···················· |
| A | 3467 | hCoV-19/Wales/PHWC-25BC8/2020\|EPI_ISL_422114\|2020- | ···················· | ···················· | ···················· | ···················· | ···················· | ···················· |
| A | 3468 | hCoV-19/Wales/PHWC-26EDD/2020\|EPI_ISL_422115\|2020- | ···················· | ···················· | ···················· | ···················· | ···················· | ···················· |
| A | 3469 | hCoV-19/Wales/PHWC-27003/2020\|EPI_ISL_422125\|2020- | ···················· | ···················· | ···················· | ···················· | ···················· | ···················· |
| A | 3470 | hCoV-19/Wales/PHWC-26699/2020\|EPI_ISL_422127\|2020- | ···················· | ···················· | ···················· | ···················· | ···················· | ···················· |
| A | 3471 | hCoV-19/Wales/PHWC-266A8/2020\|EPI_ISL_422128\|2020- | ···················· | ···················· | ···················· | ···················· | ···················· | ···················· |
| A | 3472 | hCoV-19/Wales/PHWC-267A5/2020\|EPI_ISL_422129\|2020- | ···················· | ···················· | ···················· | ···················· | ···················· | ···················· |
| A | 3473 | hCoV-19/Wales/PHWC-26A9D/2020\|EPI_ISL_422130\|2020- | ···················· | ···················· | ···················· | ···················· | ···················· | ···················· |
| A | 3474 | hCoV-19/Wales/PHWC-27030/2020\|EPI_ISL_422131\|2020- | ···················· | ···················· | ···················· | ···················· | ···················· | ···················· |
| A | 3475 | hCoV-19/Wales/PHWC-26F07/2020\|EPI_ISL_422132\|2020- | ···················· | ···················· | ···················· | ···················· | ···················· | ···················· |
| A | 3476 | hCoV-19/Wales/PHWC-270C7/2020\|EPI_ISL_422133\|2020- | ···················· | ···················· | ···················· | ···················· | ···················· | ···················· |
| A | 3477 | hCoV-19/Wales/PHWC-25E65/2020\|EPI_ISL_422135\|2020- | ···················· | ···················· | ···················· | ···················· | ···················· | ···················· |

| | | | | | | | | |
|---|---|---|---|---|---|---|---|---|
| A | 3478 hCoV-19/Wales/PHWC-25EB0/2020\|EPI_ISL_422136\|2020- | .................... | ....A............... | .................... | .................... | .................... | .................... | |
| A | 3479 hCoV-19/Wales/PHWC-26EEC/2020\|EPI_ISL_422137\|2020- | .................... | .................... | .................... | .................... | .................... | .................... | |
| A | 3480 hCoV-19/Wales/PHWC-26D94/2020\|EPI_ISL_422160\|2020- | .................... | .................... | .................... | .................... | .................... | .................... | |
| A | 3481 hCoV-19/Wales/PHWC-26CE2/2020\|EPI_ISL_422161\|2020- | .................... | .................... | .................... | .................... | .................... | .................... | |
| A | 3482 hCoV-19/Wales/PHWC-26611/2020\|EPI_ISL_422162\|2020- | .................... | .R.................. | .................... | .................... | .................... | .................... | |
| A | 3483 hCoV-19/Wales/PHWC-26D85/2020\|EPI_ISL_422163\|2020- | .................... | ....A............... | .................... | .................... | .................... | .................... | |
| A | 3484 hCoV-19/Wales/PHWC-26620/2020\|EPI_ISL_422164\|2020- | .................... | .................... | .................... | .................... | .................... | .................... | |
| A | 3485 hCoV-19/Wales/PHWC-26C3D/2020\|EPI_ISL_422165\|2020- | .................... | .................... | .................... | .................... | .................... | .................... | |
| A | 3486 hCoV-19/Wales/PHWC-26927/2020\|EPI_ISL_422166\|2020- | .................... | .................... | .................... | .................... | .................... | .................... | |
| A | 3487 hCoV-19/Wales/PHWC-266B7/2020\|EPI_ISL_422167\|2020- | .................... | .................... | .................... | .................... | .................... | .................... | |
| A | 3488 hCoV-19/Wales/PHWC-25F62/2020\|EPI_ISL_422168\|2020- | .................... | .................... | .................... | .................... | .................... | .................... | |
| A | 3489 hCoV-19/Wales/PHWC-26BA9/2020\|EPI_ISL_422170\|2020- | .................... | .................... | .................... | .................... | .................... | .................... | |
| A | 3490 hCoV-19/Wales/PHWC-26FDA/2020\|EPI_ISL_422171\|2020- | .................... | .................... | .................... | .................... | .................... | .................... | |
| A | 3491 hCoV-19/Wales/PHWC-26C4C/2020\|EPI_ISL_422173\|2020- | .................... | .................... | .................... | .................... | .................... | .................... | |
| A | 3492 hCoV-19/Wales/PHWC-26F70/2020\|EPI_ISL_422174\|2020- | .................... | .................... | .................... | .................... | .................... | .................... | |
| A | 3493 hCoV-19/Wales/PHWC-25B7D/2020\|EPI_ISL_422180\|2020- | .................... | .................... | .................... | .................... | .................... | .................... | |
| A | 3494 hCoV-19/Wales/PHWC-261F2/2020\|EPI_ISL_422181\|2020- | .................... | .................... | .................... | .................... | .................... | .................... | |
| A | 3495 hCoV-19/Wales/PHWC-267D2/2020\|EPI_ISL_422182\|2020- | .................... | .................... | .................... | .................... | .................... | .................... | |
| A | 3496 hCoV-19/Wales/PHWC-26875/2020\|EPI_ISL_422184\|2020- | .................... | .................... | .................... | .................... | .................... | .................... | |
| A | 3497 hCoV-19/Wales/PHWC-2680C/2020\|EPI_ISL_422185\|2020- | .................... | .................... | .................... | .................... | .................... | .................... | |
| A | 3498 hCoV-19/Wales/PHWC-26B4F/2020\|EPI_ISL_422186\|2020- | .................... | .................... | .................... | .................... | .................... | .................... | |
| A | 3499 hCoV-19/Wales/PHWC-2709A/2020\|EPI_ISL_422187\|2020- | .................... | .................... | .................... | .................... | .................... | .................... | |
| A | 3500 hCoV-19/Wales/PHWC-26480/2020\|EPI_ISL_422191\|2020- | .................... | .A.................. | .................... | .................... | .................... | .................... | |
| A | 3501 hCoV-19/Wales/PHWC-265BA/2020\|EPI_ISL_422192\|2020- | .................... | .................... | .................... | .................... | .................... | .................... | |
| A | 3502 hCoV-19/Wales/PHWC-26963/2020\|EPI_ISL_422193\|2020- | .................... | .................... | .................... | .................... | .................... | .................... | |
| A | 3503 hCoV-19/Wales/PHWC-2612F/2020\|EPI_ISL_422194\|2020- | .................... | .................... | .................... | .................... | .................... | .................... | |
| A | 3504 hCoV-19/Wales/PHWC-2659C/2020\|EPI_ISL_422195\|2020- | .................... | ....A............... | .................... | .................... | .................... | .................... | |
| A | 3506 hCoV-19/England/20140069404/2020\|EPI_ISL_423404\|20 | .................... | .................... | .................... | .................... | .................... | .................... | |
| A | 3509 hCoV-19/England/20140070504/2020\|EPI_ISL_423408\|20 | .................... | .................... | .................... | .................... | .................... | .................... | |
| A | 3510 hCoV-19/England/20140070604/2020\|EPI_ISL_423409\|20 | .................... | .................... | .................... | .................... | .................... | .................... | |
| A | 3511 hCoV-19/England/20140070804/2020\|EPI_ISL_423410\|20 | .................... | .................... | .................... | .................... | .................... | .................... | |
| A | 3512 hCoV-19/England/20140071104/2020\|EPI_ISL_423411\|20 | .................... | .................... | .................... | .................... | .................... | .................... | |
| A | 3514 hCoV-19/England/20140071504/2020\|EPI_ISL_423413\|20 | .................... | .................... | .................... | .................... | .................... | .................... | |
| A | 3515 hCoV-19/England/20140071604/2020\|EPI_ISL_423414\|20 | .................... | .................... | .................... | .................... | .................... | .................... | |
| A | 3516 hCoV-19/England/20140071704/2020\|EPI_ISL_423415\|20 | .................... | .................... | .................... | .................... | .................... | .................... | |
| A | 3517 hCoV-19/England/20140072304/2020\|EPI_ISL_423417\|20 | .................... | .................... | .................... | .................... | .................... | .................... | |
| A | 3518 hCoV-19/England/20140072504/2020\|EPI_ISL_423418\|20 | .................... | .................... | .................... | .................... | .................... | .................... | |
| A | 3524 hCoV-19/England/20142002704/2020\|EPI_ISL_423424\|20 | .................... | .................... | .................... | .................... | .................... | .................... | |
| A | 3525 hCoV-19/England/20142002904/2020\|EPI_ISL_423425\|20 | .................... | .................... | .................... | .................... | .................... | .................... | |
| A | 3526 hCoV-19/England/20142003904/2020\|EPI_ISL_423426\|20 | .................... | ....A............... | .................... | .................... | .................... | .................... | |
| A | 3528 hCoV-19/England/20142014504/2020\|EPI_ISL_423430\|20 | .................... | .................... | .................... | .................... | .................... | .................... | |
| A | 3529 hCoV-19/England/20142015104/2020\|EPI_ISL_423432\|20 | .................... | .................... | .................... | .................... | .................... | .................... | |
| A | 3530 hCoV-19/England/20142016204/2020\|EPI_ISL_423435\|20 | .................... | .................... | .................... | .................... | .................... | .................... | |
| A | 3531 hCoV-19/England/20142016504/2020\|EPI_ISL_423436\|20 | .................... | .................... | .................... | .................... | .................... | .................... | |
| A | 3532 hCoV-19/England/20142016704/2020\|EPI_ISL_423437\|20 | .................... | .................... | .................... | .................... | .................... | .................... | |
| A | 3533 hCoV-19/England/20142018004/2020\|EPI_ISL_423438\|20 | .................... | .................... | .................... | .................... | .................... | .................... | |
| A | 3535 hCoV-19/England/20142018804/2020\|EPI_ISL_423441\|20 | .................... | .................... | .................... | .................... | .................... | .................... | |
| A | 3538 hCoV-19/England/20142019704/2020\|EPI_ISL_423444\|20 | .................... | .................... | .................... | .................... | .................... | .NN..........N...... | 1 |
| A | 3539 hCoV-19/England/20142021604/2020\|EPI_ISL_423446\|20 | .................... | .................... | .................... | .................... | .................... | .................... | |
| A | 3542 hCoV-19/England/20142079504/2020\|EPI_ISL_423460\|20 | .................... | .................... | .................... | .................... | .................... | .................... | |
| A | 3543 hCoV-19/England/20142079804/2020\|EPI_ISL_423461\|20 | .................... | .................... | .................... | .................... | .................... | .................... | |
| A | 3545 hCoV-19/England/20142083604/2020\|EPI_ISL_423465\|20 | .................... | .................... | .................... | .................... | ..................N. | .................... | 1 |
| A | 3546 hCoV-19/England/20142083804/2020\|EPI_ISL_423466\|20 | .................... | .................... | .................... | .................... | .................... | .................... | |
| A | 3547 hCoV-19/England/20142130404/2020\|EPI_ISL_423467\|20 | .................... | .................... | .................... | .................... | .................... | .................... | |
| A | 3548 hCoV-19/England/20142130504/2020\|EPI_ISL_423468\|20 | .................... | .................... | .................... | .................... | .................... | .................... | |
| A | 3552 hCoV-19/England/20144006304/2020\|EPI_ISL_423493\|20 | .................... | .................... | .................... | .................... | .................... | .................... | |
| A | 3554 hCoV-19/England/20144006704/2020\|EPI_ISL_423495\|20 | .................... | .................... | .................... | .................... | .................... | .................... | |
| A | 3555 hCoV-19/England/20144006904/2020\|EPI_ISL_423496\|20 | .................... | .................... | .................... | .................... | .................... | .................... | |
| A | 3556 hCoV-19/England/20144007604/2020\|EPI_ISL_423497\|20 | .................... | .................... | .................... | .................... | .................... | .NN................. | |
| A | 3557 hCoV-19/England/20144008004/2020\|EPI_ISL_423498\|20 | .................... | .................... | .................... | .................... | .................... | ...N................ | |
| A | 3559 hCoV-19/USA/WA-UW-4549/2020\|EPI_ISL_429600\|2020-03 | .................... | .................... | .................... | .................... | .................... | .................... | |
| A | 3560 hCoV-19/USA/ID-UW-4552/2020\|EPI_ISL_429601\|2020-03 | .................... | .................... | .................... | .................... | .................... | .................... | |
| A | 3561 hCoV-19/USA/WA-UW-4564/2020\|EPI_ISL_429602\|2020-03 | .................... | .................... | .................... | .................... | .................... | .................... | |
| A | 3562 hCoV-19/USA/WA-UW-4565/2020\|EPI_ISL_429603\|2020-03 | .................... | .................... | .................... | .................... | .................... | .................... | |
| A | 3563 hCoV-19/USA/WA-UW-4568/2020\|EPI_ISL_429604\|2020-03 | .................... | .................... | .................... | .................... | .................... | .................... | |
| A | 3564 hCoV-19/USA/WA-UW-4578/2020\|EPI_ISL_429605\|2020-03 | .................... | .................... | .................... | .................... | .................... | .................... | |
| A | 3565 hCoV-19/USA/WA-UW-4579/2020\|EPI_ISL_429606\|2020-03 | .................... | .................... | .................... | .................... | .................... | .................... | |
| A | 3566 hCoV-19/USA/WA-UW-4593/2020\|EPI_ISL_429608\|2020-03 | .................... | .................... | .................... | .................... | .................... | .................... | |
| A | 3567 hCoV-19/USA/WA-UW-4594/2020\|EPI_ISL_429609\|2020-03 | .................... | .................... | .................... | .................... | .................... | .................... | |
| A | 3568 hCoV-19/USA/ID-UW-4598/2020\|EPI_ISL_429610\|2020-03 | .................... | .................... | .................... | .................... | .................... | .................... | |
| A | 3569 hCoV-19/USA/WA-UW-4602/2020\|EPI_ISL_429611\|2020-03 | .................... | .................... | .................... | .................... | .................... | .................... | |
| A | 3570 hCoV-19/USA/WA-UW-4609/2020\|EPI_ISL_429612\|2020-03 | .................... | .................... | .................... | .................... | .................... | .................... | |
| A | 3571 hCoV-19/USA/WA-UW-4611/2020\|EPI_ISL_429613\|2020-03 | .................... | .................... | .................... | .................... | .................... | .................... | |
| A | 3572 hCoV-19/USA/WA-UW-4612/2020\|EPI_ISL_429614\|2020-03 | .................... | .................... | .................... | .................... | .................... | .................... | |
| A | 3573 hCoV-19/USA/WA-UW-4703/2020\|EPI_ISL_429615\|2020-03 | .................... | .................... | .................... | .................... | .................... | .................... | |
| A | 3574 hCoV-19/USA/WA-UW-4732/2020\|EPI_ISL_429616\|2020-03 | .................... | .................... | .................... | .................... | .................... | .................... | |
| A | 3575 hCoV-19/USA/WA-UW-4734/2020\|EPI_ISL_429617\|2020-03 | .................... | .................... | .................... | .................... | .................... | .................... | |
| A | 3576 hCoV-19/USA/WA-UW-4741/2020\|EPI_ISL_429618\|2020-03 | .................... | .................... | .................... | .................... | .................... | .................... | |

| | | | | | | | | |
|---|---|---|---|---|---|---|---|---|
| A | 3577 | hCoV-19/USA/WA-UW-4760/2020\|EPI_ISL_429621\|2020-03 | .................... | .................... | .................... | .................... | .................... | .................... |
| A | 3578 | hCoV-19/USA/WA-UW-4763/2020\|EPI_ISL_429622\|2020-03 | .................... | .................... | .................... | .................... | .................... | .................... |
| A | 3579 | hCoV-19/USA/WA-UW-4766/2020\|EPI_ISL_429623\|2020-03 | .................... | .................... | .................... | .................... | .................... | .................... |
| A | 3580 | hCoV-19/USA/WA-UW-4769/2020\|EPI_ISL_429624\|2020-03 | .................... | .................... | .................... | .................... | .................... | .................... |
| A | 3581 | hCoV-19/USA/WA-UW-4770/2020\|EPI_ISL_429625\|2020-03 | .................... | .................... | .................... | .................... | .................... | .................... |
| A | 3582 | hCoV-19/USA/WA-UW-4775/2020\|EPI_ISL_429626\|2020-03 | .................... | .................... | .................... | .................... | .................... | .................... |
| A | 3583 | hCoV-19/USA/WA-UW-4777/2020\|EPI_ISL_429627\|2020-03 | .................... | .................... | .................... | .................... | .................... | .................... |
| A | 3584 | hCoV-19/USA/UN-UW-4645/2020\|EPI_ISL_429628\|2020-04 | .................... | .................... | .................... | .................... | .................... | .................... |
| A | 3585 | hCoV-19/Belgium/GHB-03021/2020\|EPI_ISL_407976\|2020 | .................... | .................... | .................... | .................... | .................... | .................... |
| A | 3586 | hCoV-19/Singapore/2/2020\|EPI_ISL_407987\|2020-01-25 | .................... | .................... | .................... | .................... | .................... | .................... |
| A | 3587 | hCoV-19/Singapore/3/2020\|EPI_ISL_407988\|2020-02-01 | .................... | .................... | .................... | .................... | .................... | .................... |
| A | 3588 | hCoV-19/Weifang/WF0029/2020\|EPI_ISL_413809\|2020-02 | .................... | .................... | .................... | .................... | .................... | .................... |
| A | 3589 | hCoV-19/England/20110000606/2020\|EPI_ISL_417301\|20 | .................... | .................... | .................... | .................... | .................... | .................... |
| A | 3590 | hCoV-19/England/20110000706/2020\|EPI_ISL_417302\|20 | .................... | .................... | .................... | .................... | .................... | .................... |
| A | 3591 | hCoV-19/England/20110015106/2020\|EPI_ISL_417306\|20 | .................... | .................... | .................... | .................... | .................... | .................... |
| A | 3592 | hCoV-19/England/20110023706/2020\|EPI_ISL_417307\|20 | .................... | .................... | .................... | .................... | .................... | .................... |
| A | 3593 | hCoV-19/England/20110058706/2020\|EPI_ISL_417311\|20 | .................... | .................... | .................... | .................... | .................... | .................... |
| A | 3594 | hCoV-19/England/20110059306/2020\|EPI_ISL_417312\|20 | .................... | .................... | .................... | .................... | .................... | .................... |
| A | 3595 | hCoV-19/England/20110097506/2020\|EPI_ISL_417313\|20 | .................... | .................... | .................... | .................... | .................... | .................... |
| A | 3597 | hCoV-19/England/20118044606/2020\|EPI_ISL_417315\|20 | .................... | .................... | .................... | .................... | .................... | .................... |
| A | 3598 | hCoV-19/USA/CA-SCCPHD-UC13/2020\|EPI_ISL_417317\|202 | .................... | .................... | .................... | .................... | .................... | .................... |
| A | 3599 | hCoV-19/USA/CA-SCCPHD-UC14/2020\|EPI_ISL_417318\|202 | .................... | .................... | .................... | .................... | .................... | .................... |
| A | 3600 | hCoV-19/France/ARA-06813/2020\|EPI_ISL_417340\|2020- | .................... | .................... | .................... | .................... | .................... | .................... |
| A | 3601 | hCoV-19/USA/WA-UW192/2020\|EPI_ISL_417341\|2020-03-1 | .................... | .................... | .................... | .................... | .................... | .................... |
| A | 3602 | hCoV-19/USA/WA-UW193/2020\|EPI_ISL_417342\|2020-03-1 | .................... | .................... | .................... | .................... | .................... | .................... |
| A | 3603 | hCoV-19/USA/WA-UW194/2020\|EPI_ISL_417343\|2020-03-1 | .................... | .................... | .................... | .................... | .................... | .................... |
| A | 3604 | hCoV-19/USA/WA-UW195/2020\|EPI_ISL_417344\|2020-03-1 | .................... | .................... | .................... | .................... | .................... | .................... |
| A | 3605 | hCoV-19/USA/WA-UW221/2020\|EPI_ISL_417370\|2020-03-1 | .................... | .................... | .................... | .................... | .................... | .................... |
| A | 3606 | hCoV-19/USA/WA-UW222/2020\|EPI_ISL_417371\|2020-03-1 | .................... | .................... | .................... | .................... | .................... | .................... |
| A | 3607 | hCoV-19/USA/CT-UW223/2020\|EPI_ISL_417372\|2020-03-1 | .................... | .................... | .................... | .................... | .................... | .................... |
| A | 3608 | hCoV-19/USA/CT-UW224/2020\|EPI_ISL_417373\|2020-03-1 | .................... | .................... | .................... | .................... | .................... | .................... |
| A | 3609 | hCoV-19/USA/WA-UW225/2020\|EPI_ISL_417374\|2020-03-1 | .................... | .................... | .................... | .................... | .................... | .................... |
| A | 3610 | hCoV-19/USA/WA-UW226/2020\|EPI_ISL_417375\|2020-03-1 | .................... | .................... | .................... | .................... | .................... | .................... |
| A | 3611 | hCoV-19/USA/WA-UW227/2020\|EPI_ISL_417376\|2020-03-1 | .................... | .................... | .................... | .................... | .................... | .................... |
| A | 3612 | hCoV-19/USA/WA-UW228/2020\|EPI_ISL_417377\|2020-03-1 | .................... | .................... | .................... | .................... | .................... | .................... |
| A | 3613 | hCoV-19/USA/WA-UW229/2020\|EPI_ISL_417378\|2020-03-1 | .................... | .................... | .................... | .................... | .................... | .................... |
| A | 3614 | hCoV-19/USA/WA-UW230/2020\|EPI_ISL_417379\|2020-03-1 | .................... | .................... | .................... | .................... | .................... | .................... |
| A | 3615 | hCoV-19/USA/WA-UW231/2020\|EPI_ISL_417380\|2020-03-1 | .................... | .................... | .................... | .................... | .................... | .................... |
| A | 3616 | hCoV-19/USA/WA-UW232/2020\|EPI_ISL_417381\|2020-03-1 | .................... | .................... | .................... | .................... | .................... | .................... |
| A | 3617 | hCoV-19/USA/WA-UW233/2020\|EPI_ISL_417382\|2020-03-1 | .................... | .................... | .................... | .................... | .................... | .................... |
| A | 3618 | hCoV-19/Australia/NSW15/2020\|EPI_ISL_417383\|2020-0 | .................... | .................... | .................... | .................... | .................... | .................... |
| A | 3619 | hCoV-19/Australia/NSW18/2020\|EPI_ISL_417384\|2020-0 | .................... | .................... | .................... | .................... | .................... | .................... |
| A | 3620 | hCoV-19/Australia/NSW19/2020\|EPI_ISL_417385\|2020-0 | .................... | .................... | .................... | .................... | .................... | .................... |
| A | 3621 | hCoV-19/Australia/NSW21/2020\|EPI_ISL_417386\|2020-0 | .................... | .................... | .................... | .................... | .................... | .................... |
| A | 3622 | hCoV-19/Australia/NSW24/2020\|EPI_ISL_417387\|2020-0 | .................... | .................... | .................... | .................... | .................... | .................... |
| A | 3623 | hCoV-19/Australia/NSW25/2020\|EPI_ISL_417388\|2020-0 | .................... | .................... | .................... | .................... | .................... | .................... |
| A | 3624 | hCoV-19/Australia/NSW26/2020\|EPI_ISL_417389\|2020-0 | .................... | .................... | .................... | .................... | .................... | .................... |
| A | 3625 | hCoV-19/Australia/NSW27/2020\|EPI_ISL_417390\|2020-0 | .................... | .................... | .................... | .................... | .................... | .................... |
| A | 3626 | hCoV-19/Australia/NSW28/2020\|EPI_ISL_417391\|2020-0 | .................... | .................... | .................... | .................... | .................... | .................... |
| A | 3627 | hCoV-19/Australia/NSW31/2020\|EPI_ISL_417392\|2020-0 | .................... | .................... | .................... | .................... | .................... | .................... |
| A | 3628 | hCoV-19/Australia/NSW35/2020\|EPI_ISL_417393\|2020-0 | .................... | .................... | .................... | .................... | .................... | .................... |
| A | 3629 | hCoV-19/Australia/NSW36/2020\|EPI_ISL_417394\|2020-0 | .................... | .................... | .................... | .................... | .................... | .................... |
| A | 3630 | hCoV-19/Australia/NSW37/2020\|EPI_ISL_417395\|2020-0 | .................... | .................... | .................... | .................... | .................... | .................... |
| A | 3631 | hCoV-19/Australia/NSW38/2020\|EPI_ISL_417396\|2020-0 | .................... | .................... | .................... | .................... | .................... | .................... |
| A | 3632 | hCoV-19/Australia/NSW39/2020\|EPI_ISL_417397\|2020-0 | .................... | .................... | .................... | .................... | .................... | .................... |
| A | 3633 | hCoV-19/Australia/NSW40/2020\|EPI_ISL_417398\|2020-0 | .................... | .................... | .................... | .................... | .................... | .................... |
| A | 3634 | hCoV-19/Australia/NSW41/2020\|EPI_ISL_417399\|2020-0 | .................... | .................... | .................... | .................... | .................... | .................... |
| A | 3635 | hCoV-19/Belgium/ULG-6972/2020\|EPI_ISL_418624\|2020- | .................... | .................... | .................... | .................... | .................... | .................... |
| A | 3636 | hCoV-19/Belgium/ULG-6688/2020\|EPI_ISL_418625\|2020- | .................... | .................... | .................... | .................... | .................... | .................... |
| A | 3637 | hCoV-19/Belgium/ULG-7242/2020\|EPI_ISL_418626\|2020- | .................... | ..........A......... | .................... | .................... | .................... | .................... |
| A | 3638 | hCoV-19/Belgium/ULG-6743/2020\|EPI_ISL_418627\|2020- | .................... | .................... | .................... | .................... | .................... | .................... |
| A | 3640 | hCoV-19/Belgium/ULG-7773/2020\|EPI_ISL_418629\|2020- | .................... | .................... | .................... | .................... | .................... | .................... |
| A | 3641 | hCoV-19/Belgium/ULG-7729/2020\|EPI_ISL_418630\|2020- | .................... | .................... | .................... | .................... | .................... | .................... |
| A | 3642 | hCoV-19/Belgium/ULG-6649/2020\|EPI_ISL_418631\|2020- | .................... | .................... | .................... | .................... | .................... | .................... |
| A | 3643 | hCoV-19/Belgium/ULG-7500/2020\|EPI_ISL_418632\|2020- | .................... | .................... | .................... | .................... | .................... | .................... |
| A | 3645 | hCoV-19/Belgium/ULG-8595/2020\|EPI_ISL_418634\|2020- | .................... | .................... | .................... | .................... | .................... | .................... |
| A | 3646 | hCoV-19/Belgium/ULG-8532/2020\|EPI_ISL_418635\|2020- | .................... | .................... | .................... | .................... | .................... | .................... |
| A | 3647 | hCoV-19/Belgium/ULG-8541/2020\|EPI_ISL_418636\|2020- | .................... | .................... | .................... | .................... | .................... | .................... |
| A | 3648 | hCoV-19/Belgium/ULG-8578/2020\|EPI_ISL_418638\|2020- | .................... | .................... | .................... | .................... | .................... | .................... |
| A | 3649 | hCoV-19/Belgium/ULG-8617/2020\|EPI_ISL_418639\|2020- | .................... | .................... | .................... | .................... | .................... | .................... |
| A | 3650 | hCoV-19/Belgium/ULG-8634/2020\|EPI_ISL_418640\|2020- | .................... | .................... | .................... | .................... | ..............N..... | .................... |
| A | 3651 | hCoV-19/Belgium/ULG-9558/2020\|EPI_ISL_418645\|2020- | .................... | .................... | .................... | .................... | .................... | .................... |
| A | 3652 | hCoV-19/Belgium/ULG-9572/2020\|EPI_ISL_418646\|2020- | .................... | .................... | .................... | .................... | .................... | .................... |
| A | 3653 | hCoV-19/Belgium/ULG-9618/2020\|EPI_ISL_418648\|2020- | .................... | .................... | .................... | .................... | .................... | .................... |
| A | 3655 | hCoV-19/Belgium/ULG-9620/2020\|EPI_ISL_418650\|2020- | .................... | .................... | .................... | .................... | .................... | .................... |
| A | 3656 | hCoV-19/Belgium/ULG-9634/2020\|EPI_ISL_418651\|2020- | .................... | .................... | .................... | .................... | .................... | .................... |
| A | 3657 | hCoV-19/Belgium/ULG-9641/2020\|EPI_ISL_418652\|2020- | .................... | .................... | .................... | .................... | .................... | .................... |
| A | 3658 | hCoV-19/Belgium/ULG-9647/2020\|EPI_ISL_418653\|2020- | .................... | .................... | .................... | .................... | .................... | .................... |

| | | | | | | | | |
|---|---|---|---|---|---|---|---|---|
| A | 3659 hCoV-19/Belgium/ULG-9694/2020\|EPI_ISL_418654\|2020- | ···················· | ···················· | ···················· | ···················· | ···················· | ···················· | |
| A | 3660 hCoV-19/Belgium/ULG-9714/2020\|EPI_ISL_418655\|2020- | ···················· | ···················· | ···················· | ···················· | ···················· | ···················· | |
| A | 3661 hCoV-19/Belgium/ULG-9715/2020\|EPI_ISL_418656\|2020- | ···················· | ···················· | ···················· | ···················· | ··············N····· | NN·····N············ | 1 |
| A | 3662 hCoV-19/Belgium/ULG-9716/2020\|EPI_ISL_418657\|2020- | ···················· | ···················· | ···················· | ···················· | ···················· | ···················· | |
| A | 3663 hCoV-19/Belgium/ULG-9725/2020\|EPI_ISL_418659\|2020- | ···················· | ···················· | ···················· | ···················· | ···················· | ···················· | |
| A | 3664 hCoV-19/England/20116025603/2020\|EPI_ISL_418670\|20 | ···················· | ···················· | ···················· | ···················· | ···················· | ···················· | |
| A | 3665 hCoV-19/England/20119026102/2020\|EPI_ISL_418671\|20 | ···················· | ···················· | ···················· | ···················· | ···················· | ···················· | |
| A | 3666 hCoV-19/England/20122032802/2020\|EPI_ISL_418673\|20 | ···················· | ···················· | ···················· | ···················· | ···················· | ···················· | |
| A | 3667 hCoV-19/England/20122073102/2020\|EPI_ISL_418675\|20 | ···················· | ···················· | ···················· | ···················· | ···················· | ···················· | |
| A | 3668 hCoV-19/Australia/VIC254/2020\|EPI_ISL_419949\|2020- | ···················· | ·A·················· | ···················· | ···················· | ···················· | ···················· | |
| A | 3669 hCoV-19/Australia/VIC257/2020\|EPI_ISL_419950\|2020- | ···················· | ·A·················· | ···················· | ···················· | ···················· | ···················· | |
| A | 3670 hCoV-19/Australia/VIC258/2020\|EPI_ISL_419951\|2020- | ···················· | ···················· | ···················· | ···················· | ···················· | ···················· | |
| A | 3671 hCoV-19/Australia/VIC259/2020\|EPI_ISL_419952\|2020- | ···················· | ···················· | ···················· | ···················· | ···················· | ···················· | |
| A | 3672 hCoV-19/Australia/VIC260/2020\|EPI_ISL_419953\|2020- | ···················· | ···················· | ···················· | ···················· | ···················· | ···················· | |
| A | 3673 hCoV-19/Australia/VIC261/2020\|EPI_ISL_419954\|2020- | ···················· | ···················· | ···················· | ···················· | ···················· | ···················· | |
| A | 3674 hCoV-19/Australia/VIC262/2020\|EPI_ISL_419955\|2020- | ···················· | ···················· | ···················· | ···················· | ···················· | ···················· | |
| A | 3675 hCoV-19/Australia/VIC263/2020\|EPI_ISL_419956\|2020- | ···················· | ···················· | ···················· | ···················· | ···················· | ···················· | |
| A | 3676 hCoV-19/Australia/VIC264/2020\|EPI_ISL_419957\|2020- | ···················· | ···················· | ···················· | ···················· | ······A············· | ···················· | |
| A | 3677 hCoV-19/Australia/VIC265/2020\|EPI_ISL_419958\|2020- | ···················· | ··M················· | ···················· | ···················· | ···················· | ···················· | |
| A | 3678 hCoV-19/Australia/VIC266/2020\|EPI_ISL_419959\|2020- | ···················· | ···················· | ···················· | ···················· | ······A············· | ···················· | |
| A | 3679 hCoV-19/Australia/VIC267/2020\|EPI_ISL_419960\|2020- | ···················· | ···················· | ···················· | ···················· | ···················· | ···················· | |
| A | 3680 hCoV-19/Australia/VIC268/2020\|EPI_ISL_419961\|2020- | ···················· | ···················· | ···················· | ···················· | ···················· | ···················· | |
| A | 3681 hCoV-19/Australia/VIC269/2020\|EPI_ISL_419962\|2020- | ···················· | ···················· | ···················· | ···················· | ···················· | ···················· | |
| A | 3682 hCoV-19/Australia/VIC270/2020\|EPI_ISL_419963\|2020- | ···················· | ···················· | ···················· | ···················· | ···················· | ···················· | |
| A | 3683 hCoV-19/Australia/VIC271/2020\|EPI_ISL_419964\|2020- | ···················· | ···················· | ···················· | ···················· | ···················· | ···················· | |
| A | 3684 hCoV-19/Australia/VIC272/2020\|EPI_ISL_419965\|2020- | ···················· | ···················· | ···················· | ···················· | ···················· | ···················· | |
| A | 3685 hCoV-19/Australia/VIC273/2020\|EPI_ISL_419966\|2020- | ···················· | ···················· | ···················· | ···················· | ···················· | ···················· | |
| A | 3686 hCoV-19/Australia/VIC274/2020\|EPI_ISL_419967\|2020- | ···················· | ···················· | ···················· | ···················· | ···················· | ···················· | |
| A | 3687 hCoV-19/Australia/VIC275/2020\|EPI_ISL_419968\|2020- | ···················· | ···················· | ···················· | ···················· | ···················· | ···················· | |
| A | 3688 hCoV-19/Australia/VIC276/2020\|EPI_ISL_419969\|2020- | ···················· | ···················· | ···················· | ···················· | ···················· | ···················· | |
| A | 3689 hCoV-19/Australia/VIC277/2020\|EPI_ISL_419970\|2020- | ···················· | ···················· | ···················· | ···················· | ···················· | ···················· | |
| A | 3690 hCoV-19/Australia/VIC278/2020\|EPI_ISL_419971\|2020- | ···················· | ···················· | ···················· | ···················· | ···················· | ···················· | |
| A | 3691 hCoV-19/Australia/VIC279/2020\|EPI_ISL_419972\|2020- | ···················· | ···················· | ···················· | ···················· | ···················· | ···················· | |
| A | 3692 hCoV-19/Australia/VIC280/2020\|EPI_ISL_419973\|2020- | ···················· | ···················· | ···················· | ···················· | ···················· | ···················· | |
| A | 3693 hCoV-19/Australia/VIC283/2020\|EPI_ISL_419974\|2020- | ···················· | ···················· | ···················· | ···················· | ···················· | ···················· | |
| A | 3694 hCoV-19/Australia/VIC284/2020\|EPI_ISL_419975\|2020- | ···················· | ···················· | ···················· | ···················· | ···················· | ···················· | |
| A | 3695 hCoV-19/Australia/VIC285/2020\|EPI_ISL_419976\|2020- | ···················· | ···················· | ···················· | ···················· | ···················· | ···················· | |
| A | 3696 hCoV-19/Australia/VIC286/2020\|EPI_ISL_419977\|2020- | ···················· | ···················· | ···················· | ···················· | ···················· | ···················· | |
| A | 3697 hCoV-19/Australia/VIC287/2020\|EPI_ISL_419978\|2020- | ···················· | ···················· | ···················· | ···················· | ···················· | ···················· | |
| A | 3698 hCoV-19/Australia/VIC288/2020\|EPI_ISL_419979\|2020- | ···················· | ···················· | ···················· | ···················· | ···················· | ···················· | |
| A | 3699 hCoV-19/Australia/VIC289/2020\|EPI_ISL_419980\|2020- | ···················· | ···················· | ···················· | ···················· | ···················· | ···················· | |
| A | 3700 hCoV-19/Australia/VIC291/2020\|EPI_ISL_419982\|2020- | ···················· | ···················· | ···················· | ···················· | ···················· | ···················· | |
| A | 3701 hCoV-19/Australia/VIC292/2020\|EPI_ISL_419983\|2020- | ···················· | ···················· | ···················· | ···················· | ···················· | ···················· | |
| A | 3702 hCoV-19/Australia/VIC293/2020\|EPI_ISL_419984\|2020- | ···················· | ···················· | ···················· | ···················· | ···················· | ···················· | |
| A | 3703 hCoV-19/Australia/VIC294/2020\|EPI_ISL_419985\|2020- | ···················· | ···················· | ···················· | ···················· | ···················· | ···················· | |
| A | 3704 hCoV-19/Australia/VIC295/2020\|EPI_ISL_419986\|2020- | ···················· | ···················· | ···················· | ···················· | ···················· | ···················· | |
| A | 3705 hCoV-19/Australia/VIC296/2020\|EPI_ISL_419987\|2020- | ···················· | ···················· | ···················· | ···················· | ···················· | ···················· | |
| A | 3706 hCoV-19/Australia/VIC297/2020\|EPI_ISL_419988\|2020- | ···················· | ···················· | ···················· | ···················· | ···················· | ···················· | |
| A | 3707 hCoV-19/Australia/VIC298/2020\|EPI_ISL_419989\|2020- | ···················· | ···················· | ···················· | ···················· | ···················· | ···················· | |
| A | 3708 hCoV-19/Wales/PHWC-24CB7/2020\|EPI_ISL_420937\|2020- | ···················· | ····A··············· | ···················· | ···················· | ···················· | ···················· | |
| A | 3709 hCoV-19/Wales/PHWC-24DD2/2020\|EPI_ISL_420940\|2020- | ···················· | ···················· | ···················· | ···················· | ···················· | ···················· | |
| A | 3710 hCoV-19/Wales/PHWC-24F36/2020\|EPI_ISL_420943\|2020- | ···················· | ···················· | ···················· | ···················· | ···················· | ···················· | |
| A | 3711 hCoV-19/Wales/PHWC-24FFA/2020\|EPI_ISL_420944\|2020- | ···················· | ···················· | ···················· | ···················· | ···················· | ···················· | |
| A | 3712 hCoV-19/Wales/PHWC-2522D/2020\|EPI_ISL_420949\|2020- | ···················· | ···················· | ···················· | ···················· | ···················· | ···················· | |
| A | 3713 hCoV-19/Wales/PHWC-24CF3/2020\|EPI_ISL_420951\|2020- | ···················· | ····A··············· | ···················· | ···················· | ···················· | ···················· | |
| A | 3714 hCoV-19/Wales/PHWC-24F72/2020\|EPI_ISL_420960\|2020- | ···················· | ····A··············· | ···················· | ···················· | ···················· | ···················· | |
| A | 3715 hCoV-19/Wales/PHWC-24F81/2020\|EPI_ISL_420961\|2020- | ···················· | ····A··············· | ···················· | ···················· | ···················· | ···················· | |
| A | 3716 hCoV-19/Wales/PHWC-25050/2020\|EPI_ISL_420965\|2020- | ···················· | ···················· | ···················· | ···················· | ···················· | ···················· | |
| A | 3717 hCoV-19/Wales/PHWC-25111/2020\|EPI_ISL_420969\|2020- | ···················· | ····A··············· | ···················· | ···················· | ···················· | ···················· | |
| A | 3718 hCoV-19/Wales/PHWC-25120/2020\|EPI_ISL_420970\|2020- | ···················· | ····A··············· | ···················· | ···················· | ···················· | ···················· | |
| A | 3719 hCoV-19/Wales/PHWC-25418/2020\|EPI_ISL_420990\|2020- | ···················· | ···················· | ···················· | ···················· | ···················· | ···················· | |
| A | 3720 hCoV-19/Wales/PHWC-25B04/2020\|EPI_ISL_422116\|2020- | ···················· | ···················· | ···················· | ···················· | ···················· | ···················· | |
| A | 3721 hCoV-19/Wales/PHWC-25F71/2020\|EPI_ISL_422117\|2020- | ···················· | ···················· | ···················· | ···················· | ···················· | ···················· | |
| A | 3722 hCoV-19/Wales/PHWC-25F9F/2020\|EPI_ISL_422118\|2020- | ···················· | ···················· | ···················· | ···················· | ···················· | ···················· | |
| A | 3723 hCoV-19/Wales/PHWC-26C5B/2020\|EPI_ISL_422119\|2020- | ···················· | ···················· | ···················· | ···················· | ···················· | ···················· | |
| A | 3724 hCoV-19/Wales/PHWC-26FBC/2020\|EPI_ISL_422120\|2020- | ···················· | ···················· | ···················· | ···················· | ···················· | ···················· | |
| A | 3725 hCoV-19/Wales/PHWC-26C97/2020\|EPI_ISL_422121\|2020- | ···················· | ····A··············· | ···················· | ···················· | ···················· | ···················· | |
| A | 3726 hCoV-19/Wales/PHWC-26E73/2020\|EPI_ISL_422122\|2020- | ···················· | ···················· | ···················· | ···················· | ···················· | ···················· | |
| A | 3727 hCoV-19/Wales/PHWC-26FCB/2020\|EPI_ISL_422123\|2020- | ···················· | ···················· | ···················· | ···················· | ···················· | ···················· | |
| A | 3728 hCoV-19/Wales/PHWC-2715B/2020\|EPI_ISL_422124\|2020- | ···················· | ···················· | ···················· | ···················· | ···················· | ···················· | |
| A | 3729 hCoV-19/Wales/PHWC-267B4/2020\|EPI_ISL_422126\|2020- | ···················· | ···················· | ···················· | ···················· | ···················· | ···················· | |
| A | 3730 hCoV-19/Wales/PHWC-259A0/2020\|EPI_ISL_422138\|2020- | ···················· | ···················· | ···················· | ···················· | ···················· | ···················· | |
| A | 3731 hCoV-19/Wales/PHWC-25867/2020\|EPI_ISL_422139\|2020- | ···················· | ····A··············· | ···················· | ···················· | ···················· | ···················· | |
| A | 3732 hCoV-19/Wales/PHWC-26F8F/2020\|EPI_ISL_422140\|2020- | ···················· | ···················· | ···················· | ···················· | ···················· | ···················· | |
| A | 3733 hCoV-19/Wales/PHWC-26893/2020\|EPI_ISL_422141\|2020- | ···················· | ···················· | ···················· | ···················· | ···················· | ···················· | |
| A | 3734 hCoV-19/Wales/PHWC-2613E/2020\|EPI_ISL_422142\|2020- | ···················· | ···················· | ···················· | ···················· | ···················· | ···················· | |
| A | 3735 hCoV-19/Wales/PHWC-26E64/2020\|EPI_ISL_422143\|2020- | ···················· | ···················· | ···················· | ···················· | ···················· | ···················· | |
| A | 3736 hCoV-19/Wales/PHWC-26F43/2020\|EPI_ISL_422144\|2020- | ···················· | ···················· | ···················· | ···················· | ···················· | ···················· | |

| | | | | | | | |
|---|---|---|---|---|---|---|---|
| A | 3737 hCoV-19/Wales/PHWC-26EA0/2020\|EPI_ISL_422145\|2020- | ···················· | ···················· | ···················· | ···················· | ···················· | ···················· |
| A | 3738 hCoV-19/Wales/PHWC-26D2B/2020\|EPI_ISL_422146\|2020- | ···················· | ····A··············· | ···················· | ···················· | ···················· | ···················· |
| A | 3739 hCoV-19/Wales/PHWC-2706D/2020\|EPI_ISL_422147\|2020- | ···················· | ···················· | ···················· | ···················· | ···················· | ···················· |
| A | 3740 hCoV-19/Wales/PHWC-26BC7/2020\|EPI_ISL_422148\|2020- | ···················· | ···················· | ···················· | ···················· | ···················· | ···················· |
| A | 3741 hCoV-19/Wales/PHWC-26004/2020\|EPI_ISL_422149\|2020- | ···················· | ···················· | ···················· | ···················· | ···················· | ···················· |
| A | 3742 hCoV-19/Wales/PHWC-25E0B/2020\|EPI_ISL_422150\|2020- | ···················· | ···················· | ···················· | ···················· | ···················· | ···················· |
| A | 3743 hCoV-19/Wales/PHWC-26C1F/2020\|EPI_ISL_422151\|2020- | ···················· | ···················· | ···················· | ···················· | ···················· | ···················· |
| A | 3744 hCoV-19/Wales/PHWC-26FAD/2020\|EPI_ISL_422152\|2020- | ···················· | ····A··············· | ···················· | ···················· | ···················· | ···················· |
| A | 3745 hCoV-19/Wales/PHWC-257C4/2020\|EPI_ISL_422153\|2020- | ···················· | ····A··············· | ···················· | ···················· | ···················· | ···················· |
| A | 3746 hCoV-19/Wales/PHWC-25C10/2020\|EPI_ISL_422154\|2020- | ···················· | ···················· | ···················· | ···················· | ···················· | ···················· |
| A | 3747 hCoV-19/Wales/PHWC-2713D/2020\|EPI_ISL_422155\|2020- | ···················· | ···················· | ···················· | ···················· | ···················· | ···················· |
| A | 3748 hCoV-19/Wales/PHWC-26C88/2020\|EPI_ISL_422156\|2020- | ···················· | ···················· | ···················· | ···················· | ···················· | ···················· |
| A | 3749 hCoV-19/Wales/PHWC-26787/2020\|EPI_ISL_422157\|2020- | ···················· | ···················· | ···················· | ···················· | ···················· | ···················· |
| A | 3750 hCoV-19/Wales/PHWC-266D5/2020\|EPI_ISL_422158\|2020- | ···················· | ···················· | ···················· | ···················· | ···················· | ···················· |
| A | 3751 hCoV-19/Wales/PHWC-26E28/2020\|EPI_ISL_422159\|2020- | ···················· | ···················· | ···················· | ···················· | ···················· | ···················· |
| A | 3752 hCoV-19/England/20142022104/2020\|EPI_ISL_423447\|20 | ···················· | ···················· | ···················· | ···················· | ···················· | ···················· |
| A | 3753 hCoV-19/England/20142023704/2020\|EPI_ISL_423448\|20 | ···················· | ···················· | ···················· | ···················· | ···················· | ···················· |
| A | 3758 hCoV-19/England/20142026304/2020\|EPI_ISL_423453\|20 | ···················· | ···················· | ···················· | ···················· | ···················· | ···················· |
| A | 3763 hCoV-19/England/20144000204/2020\|EPI_ISL_423472\|20 | ···················· | ···················· | ···················· | ···················· | ···················· | ···················· |
| A | 3764 hCoV-19/England/20144000404/2020\|EPI_ISL_423473\|20 | ···················· | ···················· | ···················· | ···················· | ···················· | ···················· |
| A | 3766 hCoV-19/England/20144001204/2020\|EPI_ISL_423476\|20 | ···················· | ···················· | ···················· | ···················· | ···················· | ···················· |
| A | 3768 hCoV-19/England/20144002904/2020\|EPI_ISL_423479\|20 | ···················· | ···················· | ···················· | ···················· | ···················· | ···················· |
| A | 3770 hCoV-19/England/20144003604/2020\|EPI_ISL_423481\|20 | ···················· | ···················· | ···················· | ···················· | ···················· | ···················· |
| A | 3772 hCoV-19/England/20144004504/2020\|EPI_ISL_423486\|20 | ···················· | ···················· | ···················· | ···················· | ···················· | ···················· |
| A | 3773 hCoV-19/England/20144004704/2020\|EPI_ISL_423487\|20 | ···················· | ···················· | ···················· | ···················· | ···················· | ···················· |
| A | 3774 hCoV-19/England/20144004804/2020\|EPI_ISL_423488\|20 | ···················· | ···················· | ···················· | ···················· | ···················· | ···················· |
| A | 3775 hCoV-19/Scotland/CVR69/2020\|EPI_ISL_425800\|2020-03 | ···················· | ···················· | ···················· | ···················· | ···················· | ···················· |
| A | 3776 hCoV-19/Scotland/CVR70/2020\|EPI_ISL_425801\|2020-03 | ···················· | ···················· | ···················· | ···················· | ···················· | ···················· |
| A | 3777 hCoV-19/Scotland/CVR71/2020\|EPI_ISL_425802\|2020-03 | ···················· | ···················· | ···················· | ···················· | ···················· | ···················· |
| A | 3778 hCoV-19/Scotland/CVR72/2020\|EPI_ISL_425803\|2020-03 | ···················· | ···················· | ···················· | ···················· | ···················· | ···················· |
| A | 3779 hCoV-19/Scotland/CVR74/2020\|EPI_ISL_425804\|2020-03 | ···················· | ···················· | ···················· | ···················· | ···················· | ···················· |
| A | 3780 hCoV-19/Scotland/CVR75/2020\|EPI_ISL_425805\|2020-03 | ···················· | ···················· | ···················· | ···················· | ···················· | ···················· |
| A | 3781 hCoV-19/Scotland/CVR76/2020\|EPI_ISL_425806\|2020-03 | ···················· | ···················· | ···················· | ···················· | ···················· | ···················· |
| A | 3782 hCoV-19/Scotland/CVR77/2020\|EPI_ISL_425807\|2020-03 | ···················· | ···················· | ···················· | ···················· | ···················· | ···················· |
| A | 3783 hCoV-19/Scotland/CVR78/2020\|EPI_ISL_425808\|2020-03 | ···················· | ···················· | ···················· | ···················· | ···················· | ···················· |
| A | 3784 hCoV-19/Scotland/CVR79/2020\|EPI_ISL_425809\|2020-03 | ···················· | ···················· | ···················· | ···················· | ···················· | ···················· |
| A | 3785 hCoV-19/Scotland/CVR80/2020\|EPI_ISL_425810\|2020-03 | ···················· | ···················· | ···················· | ···················· | ···················· | ···················· |
| A | 3786 hCoV-19/Scotland/CVR83/2020\|EPI_ISL_425811\|2020-03 | ···················· | ···················· | ···················· | ···················· | ···················· | ···················· |
| A | 3787 hCoV-19/Scotland/CVR85/2020\|EPI_ISL_425812\|2020-03 | ···················· | ···················· | ···················· | ···················· | ···················· | ···················· |
| A | 3788 hCoV-19/Scotland/CVR89/2020\|EPI_ISL_425813\|2020-03 | ···················· | ···················· | ···················· | ···················· | ···················· | ···················· |
| A | 3789 hCoV-19/Scotland/CVR91/2020\|EPI_ISL_425815\|2020-03 | ···················· | ·A·················· | ···················· | ···················· | ···················· | ···················· |
| A | 3790 hCoV-19/Scotland/CVR96/2020\|EPI_ISL_425816\|2020-03 | ···················· | ·A·················· | ···················· | ···················· | ···················· | ···················· |
| A | 3791 hCoV-19/Scotland/CVR97/2020\|EPI_ISL_425817\|2020-03 | ···················· | ···················· | ···················· | ···················· | ···················· | ···················· |
| A | 3792 hCoV-19/Scotland/CVR99/2020\|EPI_ISL_425818\|2020-03 | ···················· | ···················· | ···················· | ···················· | ···················· | ···················· |
| A | 3793 hCoV-19/Scotland/EDB002/2020\|EPI_ISL_425819\|2020-0 | ···················· | ···················· | ···················· | ···················· | ···················· | ···················· |
| A | 3795 hCoV-19/Scotland/EDB015/2020\|EPI_ISL_425821\|2020-0 | ···················· | ···················· | ···················· | ···················· | ···················· | ···················· |
| A | 3796 hCoV-19/Scotland/EDB016/2020\|EPI_ISL_425822\|2020-0 | ···················· | ···················· | ···················· | ···················· | ···················· | ···················· |
| A | 3797 hCoV-19/Denmark/ALAB-HH46/2020\|EPI_ISL_429300\|2020 | ···················· | ···················· | ···················· | ···················· | ···················· | ···················· |
| A | 3798 hCoV-19/Denmark/ALAB-HH49/2020\|EPI_ISL_429301\|2020 | ···················· | ···················· | ···················· | ···················· | ···················· | ···················· |
| A | 3799 hCoV-19/Denmark/ALAB-HH50/2020\|EPI_ISL_429302\|2020 | ···················· | ···················· | ···················· | ···················· | ···················· | ···················· |
| A | 3800 hCoV-19/Denmark/ALAB-HH51/2020\|EPI_ISL_429303\|2020 | ···················· | ···················· | ···················· | ···················· | ···················· | ···················· |
| A | 3801 hCoV-19/Denmark/ALAB-HH56/2020\|EPI_ISL_429304\|2020 | ···················· | ···················· | ···················· | ···················· | ···················· | ···················· |
| A | 3802 hCoV-19/Denmark/ALAB-HH57/2020\|EPI_ISL_429305\|2020 | ···················· | ···················· | ···················· | ···················· | ···················· | ···················· |
| A | 3803 hCoV-19/Denmark/ALAB-HH60/2020\|EPI_ISL_429306\|2020 | ···················· | ···················· | ···················· | ···················· | ···················· | ···················· |
| A | 3804 hCoV-19/Denmark/ALAB-HH61/2020\|EPI_ISL_429307\|2020 | ···················· | ···················· | ···················· | ···················· | ···················· | ···················· |
| A | 3805 hCoV-19/Denmark/ALAB-HH62/2020\|EPI_ISL_429308\|2020 | ···················· | ···················· | ···················· | ···················· | ···················· | ···················· |
| A | 3806 hCoV-19/Denmark/ALAB-HH63/2020\|EPI_ISL_429309\|2020 | ···················· | ···················· | ···················· | ···················· | ···················· | ···················· |
| A | 3807 hCoV-19/Denmark/ALAB-SSI121/2020\|EPI_ISL_429350\|20 | ···················· | ···················· | ···················· | ···················· | ···················· | ···················· |
| A | 3808 hCoV-19/Denmark/ALAB-SSI122/2020\|EPI_ISL_429351\|20 | ···················· | ···················· | ···················· | ···················· | ···················· | ···················· |
| A | 3809 hCoV-19/Denmark/ALAB-SSI123/2020\|EPI_ISL_429352\|20 | ···················· | ···················· | ···················· | ···················· | ···················· | ···················· |
| A | 3810 hCoV-19/Denmark/ALAB-SSI125/2020\|EPI_ISL_429353\|20 | ···················· | ···················· | ···················· | ···················· | ···················· | ···················· |
| A | 3811 hCoV-19/Denmark/ALAB-SSI126/2020\|EPI_ISL_429354\|20 | ···················· | ···················· | ···················· | ···················· | ···················· | ···················· |
| A | 3812 hCoV-19/Denmark/ALAB-SSI128/2020\|EPI_ISL_429355\|20 | ···················· | ···················· | ···················· | ···················· | ···················· | ···················· |
| A | 3813 hCoV-19/Denmark/ALAB-SSI129/2020\|EPI_ISL_429356\|20 | ···················· | ···················· | ···················· | ···················· | ···················· | ···················· |
| A | 3814 hCoV-19/Denmark/ALAB-SSI132/2020\|EPI_ISL_429357\|20 | ···················· | ···················· | ···················· | ···················· | ···················· | ···················· |
| A | 3815 hCoV-19/Denmark/ALAB-SSI139/2020\|EPI_ISL_429358\|20 | ···················· | ···················· | ···················· | ···················· | ···················· | ···················· |
| A | 3816 hCoV-19/Denmark/ALAB-SSI142/2020\|EPI_ISL_429360\|20 | ···················· | ···················· | ···················· | ···················· | ···················· | ···················· |
| A | 3817 hCoV-19/Denmark/ALAB-SSI143/2020\|EPI_ISL_429361\|20 | ···················· | ···················· | ···················· | ···················· | ···················· | ···················· |
| A | 3818 hCoV-19/Denmark/ALAB-SSI146/2020\|EPI_ISL_429364\|20 | ···················· | ···················· | ···················· | ···················· | ···················· | ···················· |
| A | 3819 hCoV-19/Denmark/ALAB-SSI150/2020\|EPI_ISL_429368\|20 | ···················· | ···················· | ···················· | ···················· | ···················· | ···················· |
| A | 3820 hCoV-19/Denmark/ALAB-SSI151/2020\|EPI_ISL_429369\|20 | ···················· | ···················· | ···················· | ···················· | ···················· | ···················· |
| A | 3821 hCoV-19/Denmark/ALAB-SSI154/2020\|EPI_ISL_429370\|20 | ···················· | ···················· | ···················· | ···················· | ···················· | ···················· |
| A | 3822 hCoV-19/Denmark/ALAB-SSI155/2020\|EPI_ISL_429371\|20 | ···················· | ···················· | ···················· | ···················· | ···················· | ···················· |
| A | 3823 hCoV-19/Denmark/ALAB-SSI156/2020\|EPI_ISL_429372\|20 | ···················· | ···················· | ···················· | ···················· | ···················· | ···················· |
| A | 3824 hCoV-19/Denmark/ALAB-SSI157/2020\|EPI_ISL_429373\|20 | ···················· | ···················· | ···················· | ···················· | ···················· | ···················· |
| A | 3825 hCoV-19/Denmark/ALAB-SSI158/2020\|EPI_ISL_429374\|20 | ···················· | ···················· | ···················· | ···················· | ···················· | ···················· |
| A | 3826 hCoV-19/Denmark/ALAB-SSI159/2020\|EPI_ISL_429375\|20 | ···················· | ···················· | ···················· | ···················· | ···················· | ···················· |
| A | 3827 hCoV-19/Denmark/ALAB-SSI160/2020\|EPI_ISL_429376\|20 | ···················· | ···················· | ···················· | ···················· | ···················· | ···················· |

A 3828 hCoV-19/Denmark/ALAB-SSI162/2020|EPI_ISL_429377|20 .................... .................... .................... .................... .................... ....................
A 3829 hCoV-19/Denmark/ALAB-SSI163/2020|EPI_ISL_429378|20 .................... .................... .................... .................... .................... ....................
A 3830 hCoV-19/Denmark/ALAB-SSI164/2020|EPI_ISL_429379|20 .................... .................... .................... .................... .................... ....................
A 3831 hCoV-19/Denmark/ALAB-SSI167/2020|EPI_ISL_429381|20 .................... .................... .................... .................... .................... ....................
A 3832 hCoV-19/Denmark/ALAB-SSI168/2020|EPI_ISL_429382|20 .................... .................... .................... .................... .................... ....................
A 3833 hCoV-19/Denmark/ALAB-SSI169/2020|EPI_ISL_429383|20 .................... .................... .................... .................... .................... ....................
A 3834 hCoV-19/Denmark/ALAB-SSI171/2020|EPI_ISL_429384|20 .................... .................... .................... .................... .................... ....................
A 3835 hCoV-19/Denmark/ALAB-SSI172/2020|EPI_ISL_429385|20 .................... .................... .................... .................... .................... ....................
A 3836 hCoV-19/Denmark/ALAB-SSI173/2020|EPI_ISL_429386|20 .................... .................... .................... .................... .................... ....................
A 3837 hCoV-19/Denmark/ALAB-SSI174/2020|EPI_ISL_429387|20 .................... .................... .................... .................... .................... ....................
A 3838 hCoV-19/Denmark/ALAB-SSI194/2020|EPI_ISL_429395|20 .................... .................... .................... .................... .................... ....................
A 3839 hCoV-19/Denmark/ALAB-SSI195/2020|EPI_ISL_429396|20 .................... .................... .................... .................... .................... ....................
A 3840 hCoV-19/Denmark/ALAB-SSI201/2020|EPI_ISL_429399|20 .................... .................... .................... .................... .................... ....................
A 3841 hCoV-19/Wales/PHW35/2020|EPI_ISL_416024|2020-03-11 .................... .................... .................... .................... .................... ....................
A 3842 hCoV-19/Wales/PHW21/2020|EPI_ISL_416026|2020-03-11 .................... .................... .................... .................... .................... ....................
A 3843 hCoV-19/Brazil/SP-07/2020|EPI_ISL_416028|2020-03-0 .................... .................... .................... .................... .................... ....................
A 3844 hCoV-19/Brazil/SP-08/2020|EPI_ISL_416029|2020-03-0 .................... .................... .................... .................... .................... ....................
A 3845 hCoV-19/Brazil/SP-09/2020|EPI_ISL_416031|2020-03-0 .................... .................... .................... .................... .................... ....................
A 3846 hCoV-19/Brazil/SP-10/2020|EPI_ISL_416032|2020-03-0 .................... .................... .................... .................... .................... ....................
A 3847 hCoV-19/Brazil/SP-11/2020|EPI_ISL_416033|2020-03-0 .................... .................... .................... .................... .................... ....................
A 3848 hCoV-19/Brazil/SP-12/2020|EPI_ISL_416034|2020-03-0 .................... .................... .................... .................... .................... ....................
A 3849 hCoV-19/Brazil/SP-13/2020|EPI_ISL_416035|2020-03-0 .................... .................... .................... .................... .................... ....................
A 3850 hCoV-19/Brazil/SP-14/2020|EPI_ISL_416036|2020-03-0 .................... .................... .................... .................... .................... ....................
A 3851 hCoV-19/USA/CA-CDPH-UC26/2020|EPI_ISL_417330|2020- .................... .................... .................... .................... .................... ....................
A 3853 hCoV-19/France/ARA-06056/2020|EPI_ISL_417334|2020- .................... .................... .................... .................... .................... ....................
A 3854 hCoV-19/France/ARA-06573/2020|EPI_ISL_417335|2020- .................... .................... .................... .................... .................... ....................
A 3855 hCoV-19/France/ARA-0668/2020|EPI_ISL_417336|2020-0 .................... .................... .................... .................... .................... ....................
A 3856 hCoV-19/France/ARA-06625/2020|EPI_ISL_417337|2020- .................... .................... .................... .................... .................... ....................
A 3857 hCoV-19/France/ARA-06756/2020|EPI_ISL_417338|2020- .................... .................... .................... .................... .................... ....................
A 3858 hCoV-19/France/ARA-06820/2020|EPI_ISL_417339|2020- .................... .................... .................... .................... .................... ....................
A 3859 hCoV-19/USA/WA-UW196/2020|EPI_ISL_417345|2020-03-1 .................... .................... .................... .................... .................... ....................
A 3860 hCoV-19/USA/WA-UW197/2020|EPI_ISL_417346|2020-03-1 .................... .................... .................... .................... .................... ....................
A 3861 hCoV-19/USA/WA-UW198/2020|EPI_ISL_417347|2020-03-1 .................... .................... .................... .................... .................... ....................
A 3862 hCoV-19/USA/WA-UW199/2020|EPI_ISL_417348|2020-03-1 .................... .................... .................... .................... .................... ....................
A 3863 hCoV-19/USA/WA-UW200/2020|EPI_ISL_417349|2020-03-1 .................... .................... .................... .................... .................... ....................
A 3864 hCoV-19/USA/WA-UW201/2020|EPI_ISL_417350|2020-03-1 .................... .................... .................... .................... .................... ....................
A 3865 hCoV-19/USA/WA-UW202/2020|EPI_ISL_417351|2020-03-1 .................... .................... .................... .................... .................... ....................
A 3866 hCoV-19/USA/WA-UW203/2020|EPI_ISL_417352|2020-03-1 .................... .................... .................... .................... .................... ....................
A 3867 hCoV-19/USA/WA-UW204/2020|EPI_ISL_417353|2020-03-1 .................... .................... .................... .................... .................... ....................
A 3868 hCoV-19/USA/WA-UW205/2020|EPI_ISL_417354|2020-03-1 .................... .................... .................... .................... .................... ....................
A 3869 hCoV-19/USA/WA-UW206/2020|EPI_ISL_417355|2020-03-1 .................... .................... .................... .................... .................... ....................
A 3870 hCoV-19/USA/WA-UW207/2020|EPI_ISL_417356|2020-03-1 .................... .................... .................... .................... .................... ....................
A 3871 hCoV-19/USA/WA-UW209/2020|EPI_ISL_417358|2020-03-1 .................... .................... .................... .................... .................... ....................
A 3872 hCoV-19/USA/WA-UW210/2020|EPI_ISL_417359|2020-03-1 .................... .................... .................... .................... .................... ....................
A 3873 hCoV-19/USA/WA-UW211/2020|EPI_ISL_417360|2020-03-1 .................... .................... .................... .................... .................... ....................
A 3874 hCoV-19/USA/WA-UW212/2020|EPI_ISL_417361|2020-03-1 .................... .................... .................... .................... .................... ....................
A 3875 hCoV-19/USA/CT-UW213/2020|EPI_ISL_417362|2020-03-1 .................... .................... .................... .................... .................... ....................
A 3876 hCoV-19/USA/WA-UW214/2020|EPI_ISL_417363|2020-03-1 .................... .................... .................... .................... .................... ....................
A 3877 hCoV-19/USA/WA-UW215/2020|EPI_ISL_417364|2020-03-1 .................... .................... .................... .................... .................... ....................
A 3878 hCoV-19/USA/WA-UW216/2020|EPI_ISL_417365|2020-03-1 .................... .................... .................... .................... .................... ....................
A 3879 hCoV-19/USA/WA-UW217/2020|EPI_ISL_417366|2020-03-1 .................... .................... .................... .................... .................... ....................
A 3880 hCoV-19/USA/WA-UW218/2020|EPI_ISL_417367|2020-03-1 .................... .................... .................... .................... .................... ....................
A 3881 hCoV-19/USA/UN-UW219/2020|EPI_ISL_417368|2020-03-1 .................... .................... .................... .................... .................... ....................
A 3882 hCoV-19/USA/WA-UW220/2020|EPI_ISL_417369|2020-03-1 .................... .................... .................... .................... .................... ....................
A 3883 hCoV-19/Belgium/ULG-9732/2020|EPI_ISL_418660|2020- .................... .................... .................... .................... .................... ....................
A 3884 hCoV-19/Belgium/ULG-9735/2020|EPI_ISL_418661|2020- .................... .................... .................... .................... .................... ....................
A 3885 hCoV-19/Belgium/ULG-9739/2020|EPI_ISL_418663|2020- .................... .................... .................... .................... .................... ....................
A 3886 hCoV-19/Belgium/ULG-9741/2020|EPI_ISL_418664|2020- .................... .................... .................... .................... .................... ....................
A 3887 hCoV-19/England/20109047803/2020|EPI_ISL_418667|20 .................... .................... .................... .................... .................... ....................
A 3888 hCoV-19/England/20110136302/2020|EPI_ISL_418668|20 .................... .................... .................... .................... .................... ....................
A 3889 hCoV-19/England/20116022902/2020|EPI_ISL_418669|20 .................... .................... .................... .................... .................... ....................
A 3890 hCoV-19/England/20120002102/2020|EPI_ISL_418672|20 .................... .................... .................... .................... .................... ....................
A 3891 hCoV-19/England/20122074602/2020|EPI_ISL_418676|20 .................... .................... .................... .................... .................... ....................
A 3892 hCoV-19/England/20122087702/2020|EPI_ISL_418677|20 .................... .................... .................... .................... .................... ....................
A 3893 hCoV-19/England/20122088102/2020|EPI_ISL_418678|20 .................... .................... .................... .................... .................... ....................
A 3894 hCoV-19/England/20122119202/2020|EPI_ISL_418679|20 .................... .................... .................... .................... .................... ....................
A 3895 hCoV-19/England/20122119302/2020|EPI_ISL_418680|20 .................... .................... .................... .................... .................... ....................
A 3896 hCoV-19/England/20122119502/2020|EPI_ISL_418681|20 .................... .................... .................... .................... .................... ....................
A 3900 hCoV-19/England/20124002002/2020|EPI_ISL_418687|20 .................... .................... .................... .................... .................... ....................
A 3902 hCoV-19/England/20124003702/2020|EPI_ISL_418690|20 .................... .................... .................... .................... .................... ....................
A 3903 hCoV-19/England/20124003902/2020|EPI_ISL_418691|20 .................... .................... .................... .................... .................... ....................
A 3904 hCoV-19/England/20124004302/2020|EPI_ISL_418692|20 .................... .................... .................... .................... .................... ....................
A 3906 hCoV-19/England/20124009602/2020|EPI_ISL_418696|20 .................... .................... .................... .................... .................... ....................
A 3908 hCoV-19/England/20124012702/2020|EPI_ISL_418698|20 .................... .................... .................... .................... .................... ....................
A 3909 hCoV-19/Australia/VIC205/2020|EPI_ISL_419900|2020- .................... .................... .................... .................... .................... ....................
A 3910 hCoV-19/Australia/VIC206/2020|EPI_ISL_419901|2020- .................... .................... .................... .................... .................... ....................
A 3911 hCoV-19/Australia/VIC207/2020|EPI_ISL_419902|2020- .................... .................... .................... .................... .................... ....................
A 3912 hCoV-19/Australia/VIC208/2020|EPI_ISL_419903|2020- .................... .................... .................... .................... .................... ....................

| | | | | | | | | |
|---|---|---|---|---|---|---|---|---|
| A | 3913 | hCoV-19/Australia/VIC209/2020\|EPI_ISL_419904\|2020- | ···················· | ···················· | ···················· | ···················· | ···················· | ···················· |
| A | 3914 | hCoV-19/Australia/VIC210/2020\|EPI_ISL_419905\|2020- | ···················· | ·A·················· | ···················· | ···················· | ···················· | ···················· |
| A | 3915 | hCoV-19/Australia/VIC211/2020\|EPI_ISL_419906\|2020- | ···················· | ···················· | ···················· | ···················· | ···················· | ···················· |
| A | 3917 | hCoV-19/Australia/VIC213/2020\|EPI_ISL_419908\|2020- | ···················· | ···················· | ···················· | ···················· | ···················· | ···················· |
| A | 3918 | hCoV-19/Australia/VIC214/2020\|EPI_ISL_419909\|2020- | ···················· | ···················· | ···················· | ···················· | ···················· | ···················· |
| A | 3919 | hCoV-19/Australia/VIC215/2020\|EPI_ISL_419910\|2020- | ···················· | ···················· | ···················· | ···················· | ···················· | ···················· |
| A | 3920 | hCoV-19/Australia/VIC216/2020\|EPI_ISL_419911\|2020- | ···················· | ···················· | ···················· | ···················· | ···················· | ···················· |
| A | 3921 | hCoV-19/Australia/VIC217/2020\|EPI_ISL_419912\|2020- | ···················· | ···················· | ···················· | ···················· | ······A············· | ···················· |
| A | 3922 | hCoV-19/Australia/VIC218/2020\|EPI_ISL_419913\|2020- | ···················· | ···················· | ···················· | ···················· | ···················· | ···················· |
| A | 3923 | hCoV-19/Australia/VIC219/2020\|EPI_ISL_419914\|2020- | ···················· | ···················· | ···················· | ···················· | ···················· | ···················· |
| A | 3924 | hCoV-19/Australia/VIC220/2020\|EPI_ISL_419915\|2020- | ···················· | ···················· | ···················· | ···················· | ···················· | ···················· |
| A | 3925 | hCoV-19/Australia/VIC221/2020\|EPI_ISL_419916\|2020- | ···················· | ···················· | ···················· | ···················· | ···················· | ···················· |
| A | 3926 | hCoV-19/Australia/VIC222/2020\|EPI_ISL_419917\|2020- | ···················· | ···················· | ···················· | ···················· | ···················· | ···················· |
| A | 3927 | hCoV-19/Australia/VIC223/2020\|EPI_ISL_419918\|2020- | ···················· | ···················· | ···················· | ···················· | ···················· | ···················· |
| A | 3928 | hCoV-19/Australia/VIC224/2020\|EPI_ISL_419919\|2020- | ···················· | ···················· | ···················· | ···················· | ······A············· | ···················· |
| A | 3929 | hCoV-19/Australia/VIC225/2020\|EPI_ISL_419920\|2020- | ···················· | ···················· | ···················· | ···················· | ···················· | ···················· |
| A | 3930 | hCoV-19/Australia/VIC226/2020\|EPI_ISL_419921\|2020- | ···················· | ···················· | ···················· | ···················· | ···················· | ···················· |
| A | 3931 | hCoV-19/Australia/VIC227/2020\|EPI_ISL_419922\|2020- | ···················· | ···················· | ···················· | ···················· | ···················· | ···················· |
| A | 3932 | hCoV-19/Australia/VIC228/2020\|EPI_ISL_419923\|2020- | ···················· | ···················· | ···················· | ···················· | ···················· | ···················· |
| A | 3933 | hCoV-19/Australia/VIC229/2020\|EPI_ISL_419924\|2020- | ···················· | ···················· | ···················· | ···················· | ···················· | ···················· |
| A | 3934 | hCoV-19/Australia/VIC230/2020\|EPI_ISL_419925\|2020- | ···················· | ···················· | ···················· | ···················· | ···················· | ···················· |
| A | 3935 | hCoV-19/Australia/VIC231/2020\|EPI_ISL_419926\|2020- | ···················· | ···················· | ···················· | ···················· | ···················· | ···················· |
| A | 3936 | hCoV-19/Australia/VIC236/2020\|EPI_ISL_419931\|2020- | ···················· | ···················· | ···················· | ···················· | ···················· | ···················· |
| A | 3937 | hCoV-19/Australia/VIC237/2020\|EPI_ISL_419932\|2020- | ···················· | ···················· | ···················· | ···················· | ···················· | ···················· |
| A | 3938 | hCoV-19/Australia/VIC238/2020\|EPI_ISL_419933\|2020- | ···················· | ···················· | ···················· | ···················· | ···················· | ···················· |
| A | 3939 | hCoV-19/Australia/VIC242/2020\|EPI_ISL_419937\|2020- | ···················· | ···················· | ···················· | ···················· | ···················· | ···················· |
| A | 3940 | hCoV-19/Australia/VIC243/2020\|EPI_ISL_419938\|2020- | ···················· | ···················· | ···················· | ···················· | ···················· | ···················· |
| A | 3941 | hCoV-19/Australia/VIC244/2020\|EPI_ISL_419939\|2020- | ···················· | ···················· | ···················· | ···················· | ···················· | ···················· |
| A | 3942 | hCoV-19/Australia/VIC245/2020\|EPI_ISL_419940\|2020- | ···················· | ···················· | ···················· | ···················· | ···················· | ···················· |
| A | 3943 | hCoV-19/Australia/VIC246/2020\|EPI_ISL_419941\|2020- | ···················· | ···················· | ···················· | ···················· | ···················· | ···················· |
| A | 3944 | hCoV-19/Australia/VIC247/2020\|EPI_ISL_419942\|2020- | ···················· | ···················· | ···················· | ···················· | ···················· | ···················· |
| A | 3945 | hCoV-19/Australia/VIC248/2020\|EPI_ISL_419943\|2020- | ···················· | ···················· | ···················· | ···················· | ···················· | ···················· |
| A | 3946 | hCoV-19/Australia/VIC249/2020\|EPI_ISL_419944\|2020- | ···················· | ···················· | ···················· | ···················· | ···················· | ···················· |
| A | 3947 | hCoV-19/Australia/VIC250/2020\|EPI_ISL_419945\|2020- | ···················· | ···················· | ···················· | ···················· | ···················· | ···················· |
| A | 3948 | hCoV-19/Australia/VIC251/2020\|EPI_ISL_419946\|2020- | ···················· | ···················· | ···················· | ···················· | ···················· | ···················· |
| A | 3949 | hCoV-19/Australia/VIC252/2020\|EPI_ISL_419947\|2020- | ···················· | ···················· | ···················· | ···················· | ···················· | ···················· |
| A | 3950 | hCoV-19/Australia/VIC253/2020\|EPI_ISL_419948\|2020- | ···················· | ···················· | ···················· | ···················· | ···················· | ···················· |
| A | 3951 | hCoV-19/Australia/VIC299/2020\|EPI_ISL_419990\|2020- | ···················· | ···················· | ···················· | ···················· | ···················· | ···················· |
| A | 3952 | hCoV-19/Australia/VIC300/2020\|EPI_ISL_419991\|2020- | ···················· | ···················· | ···················· | ···················· | ···················· | ···················· |
| A | 3953 | hCoV-19/Australia/VIC301/2020\|EPI_ISL_419992\|2020- | ···················· | ···················· | ···················· | ···················· | ···················· | ···················· |
| A | 3954 | hCoV-19/Australia/VIC302/2020\|EPI_ISL_419993\|2020- | ···················· | ···················· | ···················· | ···················· | ···················· | ···················· |
| A | 3955 | hCoV-19/Australia/VIC303/2020\|EPI_ISL_419994\|2020- | ···················· | ···················· | ···················· | ···················· | ···················· | ···················· |
| A | 3956 | hCoV-19/Australia/VIC304/2020\|EPI_ISL_419995\|2020- | ···················· | ···················· | ···················· | ···················· | ···················· | ···················· |
| A | 3957 | hCoV-19/Australia/VIC305/2020\|EPI_ISL_419996\|2020- | ···················· | ···················· | ···················· | ···················· | ···················· | ···················· |
| A | 3958 | hCoV-19/Australia/VIC306/2020\|EPI_ISL_419997\|2020- | ···················· | ···················· | ···················· | ···················· | ···················· | ···················· |
| A | 3959 | hCoV-19/Australia/VIC307/2020\|EPI_ISL_419998\|2020- | ···················· | ···················· | ···················· | ···················· | ···················· | ···················· |
| A | 3960 | hCoV-19/Australia/VIC308/2020\|EPI_ISL_419999\|2020- | ···················· | ···················· | ···················· | ···················· | ···················· | ···················· |
| A | 3962 | hCoV-19/Germany/BY-MVP-V2010840/2020\|EPI_ISL_42090 | ···················· | ···················· | ···················· | ···················· | ···················· | ···················· |
| A | 3963 | hCoV-19/Germany/BY-MVP-V2011376/2020\|EPI_ISL_42090 | ···················· | ···················· | ···················· | ···················· | ···················· | ···················· |
| A | 3964 | hCoV-19/Germany/BY-MVP-V2011395/2020\|EPI_ISL_42090 | ···················· | ···················· | ···················· | ···················· | ···················· | ···················· |
| A | 3966 | hCoV-19/Germany/BY-MVP-V2011673/2020\|EPI_ISL_42090 | ···················· | ···················· | ···················· | ···················· | ···················· | ···················· |
| A | 3967 | hCoV-19/Germany/BY-MVP-V2011675/2020\|EPI_ISL_42090 | ···················· | ···················· | ···················· | ···················· | ···················· | ···················· |
| A | 3968 | hCoV-19/Wales/PHWC-25005/2020\|EPI_ISL_420929\|2020- | ···················· | ···················· | ···················· | ···················· | ···················· | ···················· |
| A | 3969 | hCoV-19/Wales/PHWC-2516C/2020\|EPI_ISL_420974\|2020- | ···················· | ···················· | ···················· | ···················· | ···················· | ···················· |
| A | 3970 | hCoV-19/Wales/PHWC-251B7/2020\|EPI_ISL_420976\|2020- | ···················· | ···················· | ···················· | ···················· | ···················· | ···················· |
| A | 3971 | hCoV-19/Wales/PHWC-2523C/2020\|EPI_ISL_420977\|2020- | ···················· | ···················· | ···················· | ···················· | ···················· | ···················· |
| A | 3972 | hCoV-19/Wales/PHWC-253B1/2020\|EPI_ISL_420978\|2020- | ···················· | ···················· | ···················· | ···················· | ···················· | ···················· |
| A | 3973 | hCoV-19/Wales/PHWC-251D5/2020\|EPI_ISL_420982\|2020- | ···················· | ···················· | ···················· | ···················· | ···················· | ···················· |
| A | 3974 | hCoV-19/Wales/PHWC-252B4/2020\|EPI_ISL_420985\|2020- | ···················· | ···················· | ···················· | ···················· | ···················· | ···················· |
| A | 3975 | hCoV-19/Wales/PHWC-253DF/2020\|EPI_ISL_420988\|2020- | ···················· | ····A··············· | ···················· | ···················· | ···················· | ···················· |
| A | 3976 | hCoV-19/Wales/PHWC-25463/2020\|EPI_ISL_420994\|2020- | ···················· | ····A··············· | ···················· | ···················· | ···················· | ···················· |
| A | 3977 | hCoV-19/Wales/PHWC-25472/2020\|EPI_ISL_420995\|2020- | ···················· | ····A··············· | ···················· | ···················· | ···················· | ···················· |
| A | 3978 | hCoV-19/Wales/PHWC-254AF/2020\|EPI_ISL_420998\|2020- | ···················· | ···················· | ···················· | ···················· | ···················· | ···················· |
| A | 3979 | hCoV-19/England/20134025204/2020\|EPI_ISL_423129\|20 | ···················· | ···················· | ···················· | ···················· | ···················· | ···················· |
| A | 3981 | hCoV-19/England/20134025704/2020\|EPI_ISL_423131\|20 | ···················· | ···················· | ···················· | ···················· | ···················· | ···················· |
| A | 3982 | hCoV-19/England/20134025904/2020\|EPI_ISL_423132\|20 | ···················· | ···················· | ···················· | ···················· | ···················· | ···················· |
| A | 3984 | hCoV-19/England/20134027504/2020\|EPI_ISL_423136\|20 | ···················· | ···················· | ···················· | ···················· | ···················· | ···················· |
| A | 3985 | hCoV-19/England/20134027604/2020\|EPI_ISL_423137\|20 | ···················· | ···················· | ···················· | ···················· | ···················· | ···················· |
| A | 3986 | hCoV-19/England/20134027904/2020\|EPI_ISL_423138\|20 | ···················· | ···················· | ···················· | ···················· | ···················· | ···················· |
| A | 3990 | hCoV-19/England/20134060504/2020\|EPI_ISL_423147\|20 | ···················· | ···················· | ···················· | ···················· | ···················· | ···················· |
| A | 3992 | hCoV-19/England/20134060904/2020\|EPI_ISL_423149\|20 | ···················· | ···················· | ···················· | ···················· | ·N·················· | ···················· |
| A | 3993 | hCoV-19/England/20134064004/2020\|EPI_ISL_423150\|20 | ···················· | ···················· | ···················· | ···················· | ···················· | ···················· |
| A | 3994 | hCoV-19/England/20134064204/2020\|EPI_ISL_423151\|20 | ···················· | ···················· | ···················· | ···················· | ···················· | ···················· |
| A | 3995 | hCoV-19/England/20134064904/2020\|EPI_ISL_423152\|20 | ···················· | ···················· | ···················· | ···················· | ···················· | ···················· |
| A | 3996 | hCoV-19/England/20134067304/2020\|EPI_ISL_423154\|20 | ···················· | ···················· | ···················· | ···················· | ···················· | ···················· |
| A | 3998 | hCoV-19/England/20134068504/2020\|EPI_ISL_423157\|20 | ···················· | ···················· | ···················· | ···················· | ···················· | ···················· |
| A | 3999 | hCoV-19/England/20134068604/2020\|EPI_ISL_423158\|20 | ···················· | ···················· | ···················· | ···················· | ···················· | ···················· |
| A | 4001 | hCoV-19/England/20134068904/2020\|EPI_ISL_423160\|20 | ···················· | ···G················ | ···················· | ···················· | ···················· | ···················· |

| | | | | | | | | |
|---|---|---|---|---|---|---|---|---|
| A | 4003 hCoV-19/England/20134071804/2020\|EPI_ISL_423163\|20 | ···················· | ···················· | ···················· | ···················· | ···················· | ···················· | |
| A | 4006 hCoV-19/England/20134077504/2020\|EPI_ISL_423170\|20 | ···················· | ···················· | ···················· | ···················· | ···················· | ···················· | |
| A | 4007 hCoV-19/England/20134078004/2020\|EPI_ISL_423172\|20 | ···················· | ···················· | ···················· | ···················· | ···················· | ···················· | |
| A | 4009 hCoV-19/England/20134078404/2020\|EPI_ISL_423174\|20 | ···················· | ···················· | ···················· | ···················· | ···················· | ···················· | |
| A | 4011 hCoV-19/England/20134078804/2020\|EPI_ISL_423176\|20 | ···················· | ···················· | ···················· | ···················· | ···················· | ···················· | |
| A | 4013 hCoV-19/England/20134079504/2020\|EPI_ISL_423178\|20 | ···················· | ···················· | ···················· | ···················· | ···················· | ···················· | |
| A | 4014 hCoV-19/England/20134081904/2020\|EPI_ISL_423181\|20 | ···T················ | ···················· | ···················· | ···················· | ···················· | ···················· | |
| A | 4015 hCoV-19/England/20134082304/2020\|EPI_ISL_423185\|20 | ···················· | ···················· | ···················· | ···················· | ···················· | ·N·N················ | |
| A | 4017 hCoV-19/England/201360001/2020\|EPI_ISL_423194\|2020 | ···················· | ···················· | ···················· | ···················· | ···················· | ···················· | |
| A | 4018 hCoV-19/England/201360003/2020\|EPI_ISL_423195\|2020 | ···················· | ···················· | ···················· | ···················· | ···················· | ···················· | |
| A | 4019 hCoV-19/England/201360015/2020\|EPI_ISL_423196\|2020 | ···················· | ···················· | ···················· | ···················· | ···················· | ·NN················· | |
| A | 4020 hCoV-19/England/201360018/2020\|EPI_ISL_423198\|2020 | ···················· | ···················· | ···················· | ···················· | ···················· | ···················· | |
| A | 4022 hCoV-19/Iceland/435/2020\|EPI_ISL_424459\|2020-03-20 | ···················· | ···················· | ···················· | ···················· | ···················· | ···················· | |
| A | 4023 hCoV-19/Iceland/436/2020\|EPI_ISL_424460\|2020-03-20 | ···················· | ···················· | ···················· | ···················· | ···················· | ···················· | |
| A | 4024 hCoV-19/Iceland/437/2020\|EPI_ISL_424461\|2020-03-20 | ···················· | ···················· | ···················· | ···················· | ···················· | ···················· | |
| A | 4025 hCoV-19/Iceland/438/2020\|EPI_ISL_424462\|2020-03-20 | ···················· | ···················· | ···················· | ···················· | ···················· | ···················· | |
| A | 4026 hCoV-19/Iceland/439/2020\|EPI_ISL_424463\|2020-03-20 | ···················· | ···················· | ···················· | ···················· | ···················· | ···················· | |
| A | 4027 hCoV-19/Iceland/441/2020\|EPI_ISL_424465\|2020-03-20 | ···················· | ···················· | ···················· | ···················· | ···················· | ···················· | |
| A | 4028 hCoV-19/Iceland/442/2020\|EPI_ISL_424466\|2020-03-20 | ···················· | ···················· | ···················· | ···················· | ···················· | ···················· | |
| A | 4029 hCoV-19/Iceland/443/2020\|EPI_ISL_424467\|2020-03-20 | ···················· | ···················· | ···················· | ···················· | ···················· | ···················· | |
| A | 4030 hCoV-19/Iceland/444/2020\|EPI_ISL_424468\|2020-03-20 | ···················· | ···················· | ···················· | ···················· | ···················· | ···················· | |
| A | 4031 hCoV-19/Iceland/445/2020\|EPI_ISL_424469\|2020-03-20 | ···················· | ···················· | ···················· | ···················· | ···················· | ···················· | |
| A | 4032 hCoV-19/Iceland/446/2020\|EPI_ISL_424470\|2020-03-20 | ···················· | ···················· | ···················· | ···················· | ···················· | ···················· | |
| A | 4033 hCoV-19/Iceland/447/2020\|EPI_ISL_424471\|2020-03-20 | ···················· | ···················· | ···················· | ···················· | ···················· | ···················· | |
| A | 4034 hCoV-19/Iceland/448/2020\|EPI_ISL_424472\|2020-03-20 | ···················· | ···················· | ···················· | ···················· | ···················· | ···················· | |
| A | 4035 hCoV-19/Iceland/449/2020\|EPI_ISL_424473\|2020-03-20 | ···················· | ···················· | ···················· | ···················· | ···················· | ···················· | |
| A | 4036 hCoV-19/Iceland/450/2020\|EPI_ISL_424474\|2020-03-20 | ···················· | ···················· | ···················· | ···················· | ···················· | ···················· | |
| A | 4037 hCoV-19/Iceland/451/2020\|EPI_ISL_424475\|2020-03-20 | ···················· | ···················· | ···················· | ···················· | ···················· | ···················· | |
| A | 4038 hCoV-19/Iceland/452/2020\|EPI_ISL_424476\|2020-03-19 | ···················· | ···················· | ···················· | ···················· | ···················· | ···················· | |
| A | 4039 hCoV-19/Iceland/454/2020\|EPI_ISL_424477\|2020-03-20 | ···················· | ···················· | ···················· | ···················· | ···················· | ···················· | |
| A | 4040 hCoV-19/Iceland/455/2020\|EPI_ISL_424478\|2020-03-20 | ···················· | ···················· | ···················· | ···················· | ···················· | ···················· | |
| A | 4041 hCoV-19/Iceland/456/2020\|EPI_ISL_424479\|2020-03-20 | ···················· | ···················· | ···················· | ···················· | ···················· | ···················· | |
| A | 4042 hCoV-19/Iceland/457/2020\|EPI_ISL_424480\|2020-03-20 | ···················· | ···················· | ···················· | ···················· | ···················· | ···················· | |
| A | 4043 hCoV-19/Iceland/459/2020\|EPI_ISL_424482\|2020-03-20 | ···················· | ···················· | ···················· | ···················· | ···················· | ···················· | |
| A | 4044 hCoV-19/Iceland/460/2020\|EPI_ISL_424483\|2020-03-20 | ···················· | ···················· | ···················· | ···················· | ···················· | ···················· | |
| A | 4045 hCoV-19/Iceland/461/2020\|EPI_ISL_424484\|2020-03-20 | ···················· | ···················· | ···················· | ···················· | ···················· | ···················· | |
| A | 4046 hCoV-19/Iceland/462/2020\|EPI_ISL_424485\|2020-03-20 | ···················· | ···················· | ···················· | ···················· | ···················· | ···················· | |
| A | 4047 hCoV-19/Iceland/463/2020\|EPI_ISL_424486\|2020-03-20 | ···················· | ···················· | ···················· | ···················· | ···················· | ···················· | |
| A | 4048 hCoV-19/Iceland/464/2020\|EPI_ISL_424487\|2020-03-20 | ···················· | ···················· | ···················· | ···················· | ···················· | ···················· | |
| A | 4049 hCoV-19/Iceland/466/2020\|EPI_ISL_424489\|2020-03-20 | ···················· | ···················· | ···················· | ···················· | ···················· | ···················· | |
| A | 4050 hCoV-19/Iceland/467/2020\|EPI_ISL_424490\|2020-03-20 | ···················· | ···················· | ···················· | ···················· | ···················· | ···················· | |
| A | 4051 hCoV-19/Iceland/468/2020\|EPI_ISL_424491\|2020-03-20 | ···················· | ···················· | ···················· | ···················· | ···················· | ···················· | |
| A | 4052 hCoV-19/Iceland/469/2020\|EPI_ISL_424492\|2020-03-21 | ···················· | ···················· | ···················· | ···················· | ···················· | ···················· | |
| A | 4053 hCoV-19/Iceland/470/2020\|EPI_ISL_424493\|2020-03-21 | ···················· | ···················· | ···················· | ···················· | ···················· | ···················· | |
| A | 4054 hCoV-19/Iceland/471/2020\|EPI_ISL_424494\|2020-03-20 | ···················· | ···················· | ···················· | ···················· | ···················· | ···················· | |
| A | 4055 hCoV-19/Iceland/472/2020\|EPI_ISL_424495\|2020-03-20 | ···················· | ···················· | ···················· | ···················· | ···················· | ···················· | |
| A | 4056 hCoV-19/Iceland/473/2020\|EPI_ISL_424496\|2020-03-20 | ···················· | ···················· | ···················· | ···················· | ···················· | ···················· | |
| A | 4057 hCoV-19/Iceland/474/2020\|EPI_ISL_424497\|2020-03-20 | ···················· | ···················· | ···················· | ···················· | ···················· | ···················· | |
| A | 4058 hCoV-19/Iceland/475/2020\|EPI_ISL_424498\|2020-03-20 | ···················· | ···················· | ···················· | ···················· | ···················· | ···················· | |
| A | 4059 hCoV-19/Iceland/476/2020\|EPI_ISL_424499\|2020-03-20 | ···················· | ···················· | ···················· | ···················· | ···················· | ···················· | |
| A | 4060 hCoV-19/Scotland/CVR62/2020\|EPI_ISL_425790\|2020-03 | ···················· | ···················· | ···················· | ···················· | ···················· | ···················· | |
| A | 4061 hCoV-19/Scotland/CVR63/2020\|EPI_ISL_425791\|2020-03 | ···················· | ···················· | ···················· | ···················· | ···················· | ···················· | |
| A | 4062 hCoV-19/Scotland/CVR632/2020\|EPI_ISL_425792\|2020-0 | ···················· | ·A·················· | ···················· | ···················· | ···················· | ···················· | |
| A | 4063 hCoV-19/Scotland/CVR634/2020\|EPI_ISL_425793\|2020-0 | ···················· | ···················· | ···················· | ···················· | ···················· | ···················· | |
| A | 4064 hCoV-19/Scotland/CVR635/2020\|EPI_ISL_425794\|2020-0 | ···················· | ···················A | ···················· | ···················· | ···················· | ···················· | 1 |
| A | 4065 hCoV-19/Scotland/CVR64/2020\|EPI_ISL_425795\|2020-03 | ···················· | ···················· | ···················· | ···················· | ···················· | ···················· | |
| A | 4066 hCoV-19/Scotland/CVR65/2020\|EPI_ISL_425796\|2020-03 | ···················· | ···················· | ···················· | ···················· | ···················· | ···················· | |
| A | 4067 hCoV-19/Scotland/CVR66/2020\|EPI_ISL_425797\|2020-03 | ···················· | ···················· | ···················· | ···················· | ···················· | ···················· | |
| A | 4068 hCoV-19/Scotland/CVR67/2020\|EPI_ISL_425798\|2020-03 | ···················· | ···················· | ···················· | ···················· | ···················· | ···················· | |
| A | 4069 hCoV-19/Scotland/CVR68/2020\|EPI_ISL_425799\|2020-03 | ···················· | ···················· | ···················· | ···················· | ···················· | ···················· | |
| A | 4070 hCoV-19/Denmark/ALAB-HH64/2020\|EPI_ISL_429310\|2020 | ···················· | ···················· | ···················· | ···················· | ···················· | ···················· | |
| A | 4071 hCoV-19/Denmark/ALAB-HH65/2020\|EPI_ISL_429311\|2020 | ···················· | ···················· | ···················· | ···················· | ···················· | ···················· | |
| A | 4072 hCoV-19/Denmark/ALAB-HH66/2020\|EPI_ISL_429312\|2020 | ···················· | ···················· | ···················· | ···················· | ···················· | ···················· | |
| A | 4073 hCoV-19/Denmark/ALAB-HH74/2020\|EPI_ISL_429315\|2020 | ···················· | ···················· | ···················· | ···················· | ···················· | ···················· | |
| A | 4074 hCoV-19/Denmark/ALAB-HH75/2020\|EPI_ISL_429316\|2020 | ···················· | ···················· | ···················· | ···················· | ···················· | ···················· | |
| A | 4075 hCoV-19/Denmark/ALAB-HH77/2020\|EPI_ISL_429318\|2020 | ···················· | ···················· | ···················· | ···················· | ···················· | ···················· | |
| A | 4076 hCoV-19/Denmark/ALAB-HH80/2020\|EPI_ISL_429321\|2020 | ···················· | ···················· | ···················· | ···················· | ···················· | ···················· | |
| A | 4077 hCoV-19/Denmark/ALAB-HH81/2020\|EPI_ISL_429322\|2020 | ···················· | ···················· | ···················· | ···················· | ···················· | ···················· | |
| A | 4078 hCoV-19/Denmark/ALAB-HH82/2020\|EPI_ISL_429323\|2020 | ···················· | ···················· | ···················· | ···················· | ···················· | ···················· | |
| A | 4079 hCoV-19/Denmark/ALAB-HH83/2020\|EPI_ISL_429324\|2020 | ···················· | ···················· | ···················· | ···················· | ···················· | ···················· | |
| A | 4080 hCoV-19/Denmark/ALAB-HH84/2020\|EPI_ISL_429325\|2020 | ···················· | ···················· | ···················· | ···················· | ···················· | ···················· | |
| A | 4081 hCoV-19/Denmark/ALAB-HH87/2020\|EPI_ISL_429327\|2020 | ···················· | ···················· | ···················· | ···················· | ···················· | ···················· | |
| A | 4082 hCoV-19/Denmark/ALAB-HH88/2020\|EPI_ISL_429328\|2020 | ···················· | ···················· | ···················· | ···················· | ···················· | ···················· | |
| A | 4083 hCoV-19/Denmark/ALAB-HH89/2020\|EPI_ISL_429329\|2020 | ···················· | ···················· | ···················· | ···················· | ···················· | ···················· | |
| A | 4084 hCoV-19/Denmark/ALAB-HH92/2020\|EPI_ISL_429331\|2020 | ···················· | ···················· | ···················· | ···················· | ···················· | ···················· | |
| A | 4085 hCoV-19/Denmark/ALAB-SSI103/2020\|EPI_ISL_429335\|20 | ···················· | ···················· | ···················· | ···················· | ···················· | ···················· | |
| A | 4086 hCoV-19/Denmark/ALAB-SSI105/2020\|EPI_ISL_429337\|20 | ···················· | ···················· | ···················· | ···················· | ···················· | ···················· | |
| A | 4087 hCoV-19/Denmark/ALAB-SSI107/2020\|EPI_ISL_429339\|20 | ···················· | ···················· | ···················· | ···················· | ···················· | ···················· | |

| | | | | | | | |
|---|---|---|---|---|---|---|---|
| A | 4088 hCoV-19/Denmark/ALAB-SSI108/2020\|EPI_ISL_429340\|20 | ···················· | ···················· | ···················· | ···················· | ···················· | ···················· |
| A | 4089 hCoV-19/Denmark/ALAB-SSI109/2020\|EPI_ISL_429341\|20 | ···················· | ···················· | ···················· | ···················· | ···················· | ···················· |
| A | 4090 hCoV-19/Denmark/ALAB-SSI110/2020\|EPI_ISL_429342\|20 | ···················· | ···················· | ···················· | ···················· | ···················· | ···················· |
| A | 4091 hCoV-19/Denmark/ALAB-SSI111/2020\|EPI_ISL_429343\|20 | ···················· | ···················· | ···················· | ···················· | ···················· | ···················· |
| A | 4092 hCoV-19/Denmark/ALAB-SSI113/2020\|EPI_ISL_429344\|20 | ···················· | ···················· | ···················· | ···················· | ···················· | ···················· |
| A | 4093 hCoV-19/Denmark/ALAB-SSI114/2020\|EPI_ISL_429345\|20 | ···················· | ···················· | ···················· | ···················· | ···················· | ···················· |
| A | 4094 hCoV-19/Denmark/ALAB-SSI117/2020\|EPI_ISL_429346\|20 | ···················· | ···················· | ···················· | ···················· | ···················· | ···················· |
| A | 4095 hCoV-19/Denmark/ALAB-SSI119/2020\|EPI_ISL_429348\|20 | ···················· | ···················· | ···················· | ···················· | ···················· | ···················· |
| A | 4096 hCoV-19/Denmark/ALAB-SSI120/2020\|EPI_ISL_429349\|20 | ···················· | ···················· | ···················· | ···················· | ···················· | ···················· |
| A | 4097 hCoV-19/Jingzhou/HBCDC-HB-01/2020\|EPI_ISL_412459\|2 | ···················· | ···················· | ···················· | ···················· | ···················· | ···················· |
| A | 4098 hCoV-19/Weifang/WF0015/2020\|EPI_ISL_413729\|2020-02 | ···················· | ···················· | ···················· | ···················· | ···················· | ···················· |
| A | 4099 hCoV-19/Weifang/WF0016/2020\|EPI_ISL_413746\|2020-02 | ···················· | ···················· | ···················· | ···················· | ···················· | ···················· |
| A | 4100 hCoV-19/Weifang/WF0018/2020\|EPI_ISL_413748\|2020-02 | ···················· | ···················· | ···················· | ···················· | ···················· | ···················· |
| A | 4101 hCoV-19/Weifang/WF0019/2020\|EPI_ISL_413749\|2020-02 | ···················· | ···················· | ···················· | ···················· | ···················· | ···················· |
| A | 4102 hCoV-19/Weifang/WF0020/2020\|EPI_ISL_413750\|2020-02 | ···················· | ···················· | ···················· | ···················· | ···················· | ···················· |
| A | 4103 hCoV-19/Weifang/WF0021/2020\|EPI_ISL_413751\|2020-02 | ···················· | ···················· | ···················· | ···················· | ···················· | ···················· |
| A | 4104 hCoV-19/Weifang/WF0024/2020\|EPI_ISL_413753\|2020-02 | ···················· | ···················· | ···················· | ···················· | ···················· | ···················· |
| A | 4105 hCoV-19/Weifang/WF0026/2020\|EPI_ISL_413761\|2020-02 | ···················· | ···················· | ···················· | ···················· | ···················· | ···················· |
| A | 4106 hCoV-19/Weifang/WF0028/2020\|EPI_ISL_413791\|2020-02 | ···················· | ···················· | ···················· | ···················· | ···················· | ···················· |
| A | 4107 hCoV-19/USA/CA-CDPH-UC6/2020\|EPI_ISL_413924\|2020-0 | ···················· | ···················· | ···················· | ···················· | ···················· | ···················· |
| A | 4108 hCoV-19/USA/CA-CDPH-UC7/2020\|EPI_ISL_413925\|2020-0 | ···················· | ···················· | ···················· | ···················· | ···················· | ···················· |
| A | 4109 hCoV-19/USA/CA-CDPH-UC9/2020\|EPI_ISL_413928\|2020-0 | ···················· | ···················· | ···················· | ···················· | ···················· | ···················· |
| A | 4110 hCoV-19/Italy/MAR-UnivPM-1/2020\|EPI_ISL_417491\|202 | ···················· | ···················· | ···················· | ···················· | ···················· | ···················· |
| A | 4111 hCoV-19/USA/MN45-MDH45/2020\|EPI_ISL_417492\|2020-03 | ···················· | ···················· | ···················· | ···················· | ···················· | ···················· |
| A | 4112 hCoV-19/USA/MN46-MDH46/2020\|EPI_ISL_417493\|2020-03 | ···················· | ···················· | ···················· | ···················· | ···················· | ···················· |
| A | 4113 hCoV-19/USA/MN47-MDH47/2020\|EPI_ISL_417494\|2020-03 | ···················· | ···················· | ···················· | ···················· | ···················· | ···················· |
| A | 4114 hCoV-19/USA/MN48-MDH48/2020\|EPI_ISL_417495\|2020-03 | ···················· | ···················· | ···················· | ···················· | ···················· | ···················· |
| A | 4115 hCoV-19/USA/MN51-MDH51/2020\|EPI_ISL_417496\|2020-03 | ···················· | ···················· | ···················· | ···················· | ···················· | ···················· |
| A | 4116 hCoV-19/USA/MN52-MDH52/2020\|EPI_ISL_417497\|2020-03 | ···················· | ···················· | ···················· | ···················· | ···················· | ···················· |
| A | 4117 hCoV-19/USA/MN53-MDH53/2020\|EPI_ISL_417498\|2020-03 | ···················· | ···················· | ···················· | ···················· | ···················· | ···················· |
| A | 4118 hCoV-19/USA/MN55-MDH55/2020\|EPI_ISL_417499\|2020-03 | ···················· | ···················· | ···················· | ···················· | ···················· | ···················· |
| A | 4119 hCoV-19/Australia/VIC232/2020\|EPI_ISL_419927\|2020- | ···················· | ···················· | ···················· | ···················· | ···················· | ···················· |
| A | 4120 hCoV-19/Australia/VIC233/2020\|EPI_ISL_419928\|2020- | ···················· | ···················· | ···················· | ···················· | ···················· | ···················· |
| A | 4121 hCoV-19/Australia/VIC234/2020\|EPI_ISL_419929\|2020- | ···················· | ···················· | ···················· | ···················· | ···················· | ···················· |
| A | 4122 hCoV-19/Australia/VIC239/2020\|EPI_ISL_419934\|2020- | ···················· | ···················· | ···················· | ···················· | ···················· | ···················· |
| A | 4123 hCoV-19/Australia/VIC240/2020\|EPI_ISL_419935\|2020- | ···················· | ···················· | ···················· | ···················· | ···················· | ···················· |
| A | 4124 hCoV-19/Australia/VIC241/2020\|EPI_ISL_419936\|2020- | ···················· | ···················· | ···················· | ···················· | ···················· | ···················· |
| A | 4125 hCoV-19/England/20136087804/2020\|EPI_ISL_420910\|20 | ···················· | ···················· | ···················· | ···················· | ···················· | ···················· |
| A | 4126 hCoV-19/Germany/BY-MVP-V2012856/2020\|EPI_ISL_42091 | ···················· | ···················· | ···················· | ···················· | ···················· | ···················· |
| A | 4127 hCoV-19/England/20134018902/2020\|EPI_ISL_423100\|20 | ···················· | ···················· | ···················· | ···················· | ···················· | ···················· |
| A | 4128 hCoV-19/England/20134019102/2020\|EPI_ISL_423101\|20 | ···················· | ···················· | ···················· | ···················· | ···················· | N··················· |
| A | 4129 hCoV-19/England/20134019202/2020\|EPI_ISL_423102\|20 | ···················· | ···················· | ···················· | ···················· | ···················· | ···················· |
| A | 4130 hCoV-19/England/20134019304/2020\|EPI_ISL_423103\|20 | ···················· | ···················· | ···················· | ···················· | ···················· | ···················· |
| A | 4131 hCoV-19/England/20134019402/2020\|EPI_ISL_423104\|20 | ···················· | ···················· | ···················· | ···················· | ···················· | ···················· |
| A | 4132 hCoV-19/England/20134019502/2020\|EPI_ISL_423105\|20 | ···················· | ···················· | ···················· | ···················· | ···················· | ···················· |
| A | 4134 hCoV-19/England/20134020004/2020\|EPI_ISL_423108\|20 | ···················· | ···················· | ···················· | ···················· | ···················· | ····N··············· |
| A | 4135 hCoV-19/England/20134020104/2020\|EPI_ISL_423109\|20 | ···················· | ···················· | ···················· | ···················· | ···················· | ···················· |
| A | 4136 hCoV-19/England/20134020504/2020\|EPI_ISL_423110\|20 | ···················· | ···················· | ···················· | ···················· | ···················· | ···················· |
| A | 4138 hCoV-19/England/20134021004/2020\|EPI_ISL_423112\|20 | ···················· | ···················· | ···················· | ···················· | ···················· | ···················· |
| A | 4139 hCoV-19/England/20134021104/2020\|EPI_ISL_423113\|20 | ···················· | ···················· | ···················· | ···················· | ···················· | ···················· |
| A | 4140 hCoV-19/England/20134021504/2020\|EPI_ISL_423114\|20 | ···················· | ···················· | ···················· | ···················· | ···················· | ···················· |
| A | 4141 hCoV-19/England/20134021804/2020\|EPI_ISL_423117\|20 | ···················· | ···················· | ···················· | ···················· | ···················· | ···················· |
| A | 4142 hCoV-19/England/20134021904/2020\|EPI_ISL_423118\|20 | ···················· | ···················· | ···················· | ···················· | ···················· | ···················· |
| A | 4143 hCoV-19/England/20134022104/2020\|EPI_ISL_423119\|20 | ···················· | ···················· | ···················· | ···················· | ···················· | ···················· |
| A | 4145 hCoV-19/England/20134023104/2020\|EPI_ISL_423122\|20 | ···················· | ···················· | ···················· | ···················· | ···················· | ···················· |
| A | 4146 hCoV-19/England/20134023204/2020\|EPI_ISL_423123\|20 | ···················· | ···················· | ···················· | ···················· | ···················· | ···················· |
| A | 4148 hCoV-19/England/20134024104/2020\|EPI_ISL_423126\|20 | ···················· | ···················· | ···················· | ···················· | ···················· | ···················· |
| A | 4149 hCoV-19/England/201340860/2020\|EPI_ISL_423192\|2020 | ···················· | ···················· | ···················· | ···················· | ···················· | ···················· |
| A | 4150 hCoV-19/Iceland/376/2020\|EPI_ISL_424400\|2020-03-19 | ···················· | ···················· | ···················· | ···················· | ···················· | ···················· |
| A | 4151 hCoV-19/Iceland/377/2020\|EPI_ISL_424401\|2020-03-19 | ···················· | ···················· | ···················· | ···················· | ···················· | ···················· |
| A | 4152 hCoV-19/Iceland/378/2020\|EPI_ISL_424402\|2020-03-19 | ···················· | ···················· | ···················· | ···················· | ···················· | ···················· |
| A | 4153 hCoV-19/Iceland/379/2020\|EPI_ISL_424403\|2020-03-18 | ···················· | ···················· | ···················· | ···················· | ···················· | ···················· |
| A | 4154 hCoV-19/Iceland/380/2020\|EPI_ISL_424404\|2020-03-19 | ···················· | ···················· | ···················· | ···················· | ···················· | ···················· |
| A | 4155 hCoV-19/Iceland/381/2020\|EPI_ISL_424405\|2020-03-19 | ···················· | ···················· | ···················· | ···················· | ···················· | ···················· |
| A | 4156 hCoV-19/Iceland/382/2020\|EPI_ISL_424406\|2020-03-19 | ···················· | ···················· | ···················· | ···················· | ···················· | ···················· |
| A | 4157 hCoV-19/Iceland/383/2020\|EPI_ISL_424407\|2020-03-19 | ···················· | ···················· | ···················· | ···················· | ···················· | ···················· |
| A | 4158 hCoV-19/Iceland/385/2020\|EPI_ISL_424409\|2020-03-19 | ···················· | ···················· | ···················· | ···················· | ···················· | ···················· |
| A | 4159 hCoV-19/Iceland/386/2020\|EPI_ISL_424410\|2020-03-19 | ···················· | ···················· | ···················· | ···················· | ···················· | ···················· |
| A | 4160 hCoV-19/Iceland/387/2020\|EPI_ISL_424411\|2020-03-19 | ···················· | ···················· | ···················· | ···················· | ···················· | ···················· |
| A | 4161 hCoV-19/Iceland/388/2020\|EPI_ISL_424412\|2020-03-19 | ···················· | ···················· | ···················· | ···················· | ···················· | ···················· |
| A | 4162 hCoV-19/Iceland/389/2020\|EPI_ISL_424413\|2020-03-19 | ···················· | ···················· | ···················· | ···················· | ···················· | ···················· |
| A | 4163 hCoV-19/Iceland/390/2020\|EPI_ISL_424414\|2020-03-19 | ···················· | ···················· | ···················· | ···················· | ···················· | ···················· |
| A | 4164 hCoV-19/Iceland/391/2020\|EPI_ISL_424415\|2020-03-19 | ···················· | ···················· | ···················· | ···················· | ···················· | ···················· |
| A | 4165 hCoV-19/Iceland/392/2020\|EPI_ISL_424416\|2020-03-19 | ···················· | ···················· | ···················· | ···················· | ···················· | ···················· |
| A | 4166 hCoV-19/Iceland/393/2020\|EPI_ISL_424417\|2020-03-19 | ···················· | ···················· | ···················· | ···················· | ···················· | ···················· |
| A | 4167 hCoV-19/Iceland/394/2020\|EPI_ISL_424418\|2020-03-19 | ···················· | ···················· | ···················· | ···················· | ···················· | ···················· |
| A | 4168 hCoV-19/Iceland/395/2020\|EPI_ISL_424419\|2020-03-19 | ···················· | ···················· | ···················· | ···················· | ···················· | ···················· |
| A | 4169 hCoV-19/Iceland/396/2020\|EPI_ISL_424420\|2020-03-19 | ···················· | ···················· | ···················· | ···················· | ···················· | ···················· |

| | | | | | | | | | |
|---|---|---|---|---|---|---|---|---|---|
| A | 4170 | hCoV-19/Iceland/398/2020\|EPI_ISL_424422\|2020-03-19 | ···················· | ···················· | ····················· | ···················· | ···················· | ···················· | |
| A | 4171 | hCoV-19/Iceland/399/2020\|EPI_ISL_424423\|2020-03-19 | ···················· | ···················· | ····················· | ···················· | ···················· | ···················· | |
| A | 4172 | hCoV-19/Iceland/400/2020\|EPI_ISL_424424\|2020-03-19 | ···················· | ···················· | ····················· | ···················· | ···················· | ···················· | |
| A | 4173 | hCoV-19/Iceland/401/2020\|EPI_ISL_424425\|2020-03-19 | ···················· | ···················· | ····················· | ···················· | ···················· | ···················· | |
| A | 4174 | hCoV-19/Iceland/402/2020\|EPI_ISL_424426\|2020-03-19 | ···················· | ···················· | ····················· | ···················· | ···················· | ···················· | |
| A | 4175 | hCoV-19/Iceland/403/2020\|EPI_ISL_424427\|2020-03-19 | ···················· | ···················· | ····················· | ···················· | ···················· | ···················· | |
| A | 4176 | hCoV-19/Iceland/404/2020\|EPI_ISL_424428\|2020-03-19 | ···T················ | ···················· | ····················· | ···················· | ···················· | ···················· | |
| A | 4177 | hCoV-19/Iceland/405/2020\|EPI_ISL_424429\|2020-03-19 | ···················· | ···················· | ····················· | ···················· | ···················· | ···················· | |
| A | 4178 | hCoV-19/Iceland/406/2020\|EPI_ISL_424430\|2020-03-19 | ···················· | ···················· | ····················· | ···················· | ···················· | ···················· | |
| A | 4179 | hCoV-19/Iceland/407/2020\|EPI_ISL_424431\|2020-03-19 | ···················· | ···················· | ····················· | ···················· | ···················· | ···················· | |
| A | 4180 | hCoV-19/Iceland/408/2020\|EPI_ISL_424432\|2020-03-19 | ···················· | ···················· | ····················· | ···················· | ···················· | ···················· | |
| A | 4181 | hCoV-19/Iceland/409/2020\|EPI_ISL_424433\|2020-03-19 | ···················· | ···················· | ····················· | ···················· | ···················· | ···················· | |
| A | 4182 | hCoV-19/Iceland/410/2020\|EPI_ISL_424434\|2020-03-19 | ···················· | ···················· | ····················· | ···················· | ···················· | ···················· | |
| A | 4183 | hCoV-19/Iceland/411/2020\|EPI_ISL_424435\|2020-03-19 | ···················· | ···················· | ····················· | ···················· | ···················· | ···················· | |
| A | 4184 | hCoV-19/Iceland/412/2020\|EPI_ISL_424436\|2020-03-19 | ···················· | ···················· | ····················· | ···················· | ···················· | ···················· | |
| A | 4185 | hCoV-19/Iceland/413/2020\|EPI_ISL_424437\|2020-03-19 | ···················· | ···················· | ····················· | ···················· | ···················· | ···················· | |
| A | 4186 | hCoV-19/Iceland/414/2020\|EPI_ISL_424438\|2020-03-19 | ···················· | ···················· | ····················· | ···················· | ···················· | ···················· | |
| A | 4187 | hCoV-19/Iceland/415/2020\|EPI_ISL_424439\|2020-03-19 | ···················· | ···················· | ····················· | ···················· | ···················· | ···················· | |
| A | 4188 | hCoV-19/Iceland/416/2020\|EPI_ISL_424440\|2020-03-19 | ···················· | ···················· | ····················· | ···················· | ···················· | ···················· | |
| A | 4189 | hCoV-19/Iceland/417/2020\|EPI_ISL_424441\|2020-03-19 | ···················· | ···················· | ····················· | ···················· | ···················· | ···················· | |
| A | 4190 | hCoV-19/Iceland/418/2020\|EPI_ISL_424442\|2020-03-19 | ···················· | ···················A | ····················· | ···················· | ···················· | ···················· | 1 |
| A | 4191 | hCoV-19/Iceland/419/2020\|EPI_ISL_424443\|2020-03-19 | ···················· | ···················· | ····················· | ···················· | ···················· | ···················· | |
| A | 4192 | hCoV-19/Iceland/420/2020\|EPI_ISL_424444\|2020-03-19 | ···················· | ···················· | ····················· | ···················· | ···················· | ···················· | |
| A | 4193 | hCoV-19/Iceland/421/2020\|EPI_ISL_424445\|2020-03-19 | ···················· | ···················· | ····················· | ···················· | ···················· | ···················· | |
| A | 4194 | hCoV-19/Iceland/422/2020\|EPI_ISL_424446\|2020-03-19 | ···················· | ···················· | ····················· | ···················· | ···················· | ···················· | |
| A | 4195 | hCoV-19/Iceland/423/2020\|EPI_ISL_424447\|2020-03-19 | ···················· | ···················· | ····················· | ···················· | ···················· | ···················· | |
| A | 4196 | hCoV-19/Iceland/424/2020\|EPI_ISL_424448\|2020-03-20 | ···················· | ···················· | ····················· | ···················· | ···················· | ···················· | |
| A | 4197 | hCoV-19/Iceland/425/2020\|EPI_ISL_424449\|2020-03-20 | ···················· | ···················· | ····················· | ···················· | ···················· | ···················· | |
| A | 4198 | hCoV-19/Iceland/426/2020\|EPI_ISL_424450\|2020-03-20 | ···················· | ···················· | ····················· | ···················· | ···················· | ···················· | |
| A | 4199 | hCoV-19/Iceland/427/2020\|EPI_ISL_424451\|2020-03-20 | ···················· | ···················· | ····················· | ···················· | ···················· | ···················· | |
| A | 4200 | hCoV-19/Iceland/428/2020\|EPI_ISL_424452\|2020-03-20 | ···················· | ···················· | ····················· | ···················· | ···················· | ···················· | |
| A | 4201 | hCoV-19/Iceland/429/2020\|EPI_ISL_424453\|2020-03-20 | ···················· | ···················· | ····················· | ···················· | ···················· | ···················· | |
| A | 4202 | hCoV-19/Iceland/430/2020\|EPI_ISL_424454\|2020-03-20 | ···················· | ···················· | ····················· | ···················· | ···················· | ···················· | |
| A | 4203 | hCoV-19/Iceland/431/2020\|EPI_ISL_424455\|2020-03-20 | ···················· | ···················· | ····················· | ···················· | ···················· | ···················· | |
| A | 4204 | hCoV-19/Iceland/432/2020\|EPI_ISL_424456\|2020-03-20 | ···················· | ···················· | ····················· | ···················· | ···················· | ···················· | |
| A | 4205 | hCoV-19/Iceland/433/2020\|EPI_ISL_424457\|2020-03-20 | ···················· | ···················· | ····················· | ···················· | ···················· | ···················· | |
| A | 4206 | hCoV-19/Iceland/434/2020\|EPI_ISL_424458\|2020-03-20 | ···················· | ···················· | ····················· | ···················· | ···················· | ···················· | |
| A | 4207 | hCoV-19/Iceland/581/2020\|EPI_ISL_424601\|2020-03-27 | ···················· | ···················· | ····················· | ···················· | ···················· | ···················· | |
| A | 4208 | hCoV-19/Iceland/582/2020\|EPI_ISL_424602\|2020-03-27 | ···················· | ···················· | ····················· | ···················· | ···················· | ···················· | |
| A | 4209 | hCoV-19/Iceland/583/2020\|EPI_ISL_424603\|2020-03-27 | ···················· | ···················· | ····················· | ···················· | ···················· | ···················· | |
| A | 4210 | hCoV-19/Iceland/584/2020\|EPI_ISL_424604\|2020-03-27 | ···················· | ···················· | ····················· | ···················· | ···················· | ···················· | |
| A | 4211 | hCoV-19/Iceland/585/2020\|EPI_ISL_424605\|2020-03-27 | ···················· | ···················· | ····················· | ···················· | ···················· | ···················· | |
| A | 4212 | hCoV-19/Iceland/586/2020\|EPI_ISL_424606\|2020-03-28 | ···················· | ···················· | ····················· | ···················· | ···················· | ···················· | |
| A | 4213 | hCoV-19/Iceland/588/2020\|EPI_ISL_424608\|2020-03-29 | ···················· | ···················· | ····················· | ···················· | ···················· | ···················· | |
| A | 4214 | hCoV-19/Iceland/589/2020\|EPI_ISL_424609\|2020-03-29 | ···················· | ···················· | ····················· | ···················· | ···················· | ···················· | |
| A | 4215 | hCoV-19/Iceland/590/2020\|EPI_ISL_424610\|2020-03-26 | ···················· | ···················· | ····················· | ···················· | ···················· | ···················· | |
| A | 4216 | hCoV-19/Iceland/592/2020\|EPI_ISL_424612\|2020-03-27 | ···················· | ···················· | ····················· | ···················· | ···················· | ···················· | |
| A | 4217 | hCoV-19/Scotland/CVR163/2020\|EPI_ISL_425702\|2020-0 | ···················· | ···················· | ····················· | ···················· | ···················· | ···················· | |
| A | 4218 | hCoV-19/Scotland/CVR164/2020\|EPI_ISL_425703\|2020-0 | ···················· | ···················· | ····················· | ···················· | ···················· | ···················· | |
| A | 4219 | hCoV-19/Scotland/CVR165/2020\|EPI_ISL_425704\|2020-0 | ···················· | ···················· | ····················· | ···················· | ···················· | ···················· | |
| A | 4220 | hCoV-19/Scotland/CVR166/2020\|EPI_ISL_425705\|2020-0 | ···················· | ···················· | ····················· | ···················· | ···················· | ···················· | |
| A | 4221 | hCoV-19/Scotland/CVR168/2020\|EPI_ISL_425707\|2020-0 | ···················· | ···················· | ····················· | ···················· | ···················· | ···················· | |
| A | 4222 | hCoV-19/Scotland/CVR169/2020\|EPI_ISL_425708\|2020-0 | ···················· | ···················· | ····················· | ···················· | ···················· | ···················· | |
| A | 4223 | hCoV-19/Scotland/CVR17/2020\|EPI_ISL_425709\|2020-03 | ···················· | ···················· | ····················· | ···················· | ···················· | ···················· | |
| A | 4224 | hCoV-19/Scotland/CVR171/2020\|EPI_ISL_425711\|2020-0 | ···················· | ···················· | ····················· | ···················· | ···················· | ···················· | |
| A | 4225 | hCoV-19/Scotland/CVR173/2020\|EPI_ISL_425713\|2020-0 | ···················· | ···················· | ····················· | ···················· | ···················· | ···················· | |
| A | 4226 | hCoV-19/Scotland/CVR175/2020\|EPI_ISL_425715\|2020-0 | ···················· | ···················· | ····················· | ···················· | ···················· | ···················· | |
| A | 4227 | hCoV-19/Scotland/CVR176/2020\|EPI_ISL_425716\|2020-0 | ···················· | ···················· | ····················· | ···················· | ···················· | ···················· | |
| A | 4228 | hCoV-19/Scotland/CVR177/2020\|EPI_ISL_425717\|2020-0 | ···················· | ···················· | ····················· | ···················· | ···················· | ···················· | |
| A | 4229 | hCoV-19/Scotland/CVR18/2020\|EPI_ISL_425720\|2020-03 | ···················· | ···················· | ····················· | ···················· | ···················· | ···················· | |
| A | 4230 | hCoV-19/Scotland/CVR180/2020\|EPI_ISL_425721\|2020-0 | ···················· | ···················· | ····················· | ···················· | ···················· | ···················· | |
| A | 4231 | hCoV-19/Scotland/CVR183/2020\|EPI_ISL_425722\|2020-0 | ···················· | ···················· | ····················· | ···················· | ···················· | ···················· | |
| A | 4232 | hCoV-19/Scotland/CVR19/2020\|EPI_ISL_425724\|2020-03 | ···················· | ···················· | ····················· | ···················· | ···················· | ···················· | |
| A | 4233 | hCoV-19/Scotland/CVR190/2020\|EPI_ISL_425725\|2020-0 | ···················· | ···················· | ····················· | ···················· | ···················· | ···················· | |
| A | 4234 | hCoV-19/Scotland/CVR191/2020\|EPI_ISL_425726\|2020-0 | ···················· | ···················· | ····················· | ···················· | ···················· | ···················· | |
| A | 4235 | hCoV-19/Scotland/CVR195/2020\|EPI_ISL_425727\|2020-0 | ···················· | ···················· | ····················· | ···················· | ···················· | ···················· | |
| A | 4236 | hCoV-19/Scotland/CVR196/2020\|EPI_ISL_425728\|2020-0 | ···················· | ·A·················· | ····················· | ···················· | ···················· | ···················· | |
| A | 4237 | hCoV-19/Scotland/CVR199/2020\|EPI_ISL_425729\|2020-0 | ···················· | ···················· | ····················· | ···················· | ···················· | ···················· | |
| A | 4238 | hCoV-19/Scotland/CVR20/2020\|EPI_ISL_425730\|2020-03 | ···················· | ···················· | ····················· | ···················· | ···················· | ···················· | |
| A | 4239 | hCoV-19/Scotland/CVR200/2020\|EPI_ISL_425731\|2020-0 | ···················· | ···················· | ····················· | ···················· | ···················· | ···················· | |
| A | 4240 | hCoV-19/Scotland/CVR201/2020\|EPI_ISL_425732\|2020-0 | ···················· | ···················· | ····················· | ···················· | ···················· | ···················· | |
| A | 4241 | hCoV-19/Scotland/CVR202/2020\|EPI_ISL_425733\|2020-0 | ···················· | ···················· | ····················· | ···················· | ···················· | ···················· | |
| A | 4242 | hCoV-19/Scotland/CVR203/2020\|EPI_ISL_425734\|2020-0 | ···················· | ···················· | ····················· | ···················· | ···················· | ···················· | |
| A | 4243 | hCoV-19/Scotland/CVR205/2020\|EPI_ISL_425735\|2020-0 | ···················· | ···················· | ····················· | ···················· | ···················· | ···················· | |
| A | 4244 | hCoV-19/Scotland/CVR206/2020\|EPI_ISL_425736\|2020-0 | ···················· | ···················· | ····················· | ···················· | ···················· | ···················· | |
| A | 4245 | hCoV-19/Scotland/CVR207/2020\|EPI_ISL_425737\|2020-0 | ···················· | ···················· | ····················· | ···················· | ···················· | ···················· | |
| A | 4246 | hCoV-19/Scotland/CVR21/2020\|EPI_ISL_425739\|2020-03 | ···················· | ···················· | ····················· | ···················· | ···················· | ···················· | |
| A | 4247 | hCoV-19/Scotland/CVR211/2020\|EPI_ISL_425740\|2020-0 | ···················· | ···················· | ····················· | ···················· | ···················· | ···················· | |

| | | | | | | | | |
|---|---|---|---|---|---|---|---|---|
| A | 4248 hCoV-19/Scotland/CVR212/2020\|EPI_ISL_425741\|2020-0 | ···················· | ···················· | ···················· | ···················· | ···················· | ···················· | |
| A | 4249 hCoV-19/Scotland/CVR213/2020\|EPI_ISL_425742\|2020-0 | ···················· | ···················· | ···················· | ···················· | ···················· | ···················· | |
| A | 4250 hCoV-19/Scotland/CVR215/2020\|EPI_ISL_425743\|2020-0 | ···················· | ···················· | ···················· | ···················· | ···················· | ···················· | |
| A | 4251 hCoV-19/Scotland/CVR216/2020\|EPI_ISL_425744\|2020-0 | ···················· | ···················· | ···················· | ···················· | ···················· | ···················· | |
| A | 4252 hCoV-19/Scotland/CVR218/2020\|EPI_ISL_425745\|2020-0 | ···················· | ·A·················· | ···················· | ···················· | ···················· | ···················· | |
| A | 4253 hCoV-19/Scotland/CVR219/2020\|EPI_ISL_425746\|2020-0 | ···················· | ···················· | ···················· | ···················· | ···················· | ···················· | |
| A | 4254 hCoV-19/Scotland/CVR220/2020\|EPI_ISL_425747\|2020-0 | ···················· | ···················· | ···················· | ···················· | ···················· | ···················· | |
| A | 4255 hCoV-19/Scotland/CVR222/2020\|EPI_ISL_425748\|2020-0 | ···················· | ···················· | ···················· | ···················· | ···················· | ···················· | |
| A | 4256 hCoV-19/Scotland/CVR23/2020\|EPI_ISL_425749\|2020-03 | ···················· | ···················· | ···················· | ···················· | ···················· | ···················· | |
| A | 4257 hCoV-19/Scotland/CVR235/2020\|EPI_ISL_425750\|2020-0 | ···················· | ···················· | ···················· | ···················· | ···················· | ···················· | |
| A | 4258 hCoV-19/Scotland/CVR236/2020\|EPI_ISL_425751\|2020-0 | ···················· | ···················· | ···················· | ···················· | ···················· | ···················· | |
| A | 4259 hCoV-19/Scotland/CVR24/2020\|EPI_ISL_425752\|2020-03 | ···················· | ···················· | ···················· | ···················· | ···················· | ···················· | |
| A | 4260 hCoV-19/Scotland/CVR264/2020\|EPI_ISL_425754\|2020-0 | ···················· | ···················· | ···················· | ···················· | ···················· | ···················· | |
| A | 4261 hCoV-19/Scotland/CVR268/2020\|EPI_ISL_425755\|2020-0 | ···················· | ···················· | ···················· | ···················· | ···················· | ···················· | |
| A | 4262 hCoV-19/Scotland/CVR27/2020\|EPI_ISL_425756\|2020-03 | ···················· | ···················· | ···················· | ···················· | ···················· | ···················· | |
| A | 4263 hCoV-19/Scotland/CVR28/2020\|EPI_ISL_425757\|2020-03 | ···················· | ···················· | ···················· | ···················· | ···················· | ···················· | |
| A | 4264 hCoV-19/Scotland/CVR29/2020\|EPI_ISL_425758\|2020-03 | ···················· | ···················· | ···················· | ···················· | ···················· | ···················· | |
| A | 4265 hCoV-19/Scotland/CVR30/2020\|EPI_ISL_425759\|2020-03 | ···················· | ···················· | ···················· | ···················· | ···················· | ···················· | |
| A | 4266 hCoV-19/Scotland/CVR31/2020\|EPI_ISL_425760\|2020-03 | ···················· | ···················· | ···················· | ···················· | ···················· | ···················· | |
| A | 4267 hCoV-19/Scotland/CVR32/2020\|EPI_ISL_425761\|2020-03 | ···················· | ···················· | ···················· | ···················· | ···················· | ···················· | |
| A | 4268 hCoV-19/Scotland/CVR33/2020\|EPI_ISL_425762\|2020-03 | ···················· | ·A·················· | ···················· | ···················· | ···················· | ···················· | |
| A | 4269 hCoV-19/Scotland/CVR34/2020\|EPI_ISL_425763\|2020-03 | ···················· | ···················· | ···················· | ···················· | ···················· | ···················· | |
| A | 4270 hCoV-19/Scotland/CVR35/2020\|EPI_ISL_425764\|2020-03 | ···················· | ···················· | ···················· | ···················· | ···················· | ···················· | |
| A | 4271 hCoV-19/Scotland/CVR36/2020\|EPI_ISL_425765\|2020-03 | ···················· | ···················· | ···················· | ···················· | ···················· | ···················· | |
| A | 4272 hCoV-19/Scotland/CVR37/2020\|EPI_ISL_425766\|2020-03 | ···················· | ···················· | ···················· | ···················· | ···················· | ···················· | |
| A | 4273 hCoV-19/Scotland/CVR38/2020\|EPI_ISL_425767\|2020-03 | ···················· | ···················· | ···················· | ···················· | ···················· | ···················· | |
| A | 4274 hCoV-19/Scotland/CVR40/2020\|EPI_ISL_425768\|2020-03 | ···················· | ···················· | ···················· | ···················· | ···················· | ···················· | |
| A | 4275 hCoV-19/Scotland/CVR41/2020\|EPI_ISL_425769\|2020-03 | ···················· | ···················· | ···················· | ···················· | ···················· | ···················· | |
| A | 4276 hCoV-19/Scotland/CVR42/2020\|EPI_ISL_425770\|2020-03 | ···················· | ···················· | ···················· | ···················· | ···················· | ···················· | |
| A | 4277 hCoV-19/Scotland/CVR44/2020\|EPI_ISL_425772\|2020-03 | ···················· | ···················· | ···················· | ···················· | ···················· | ···················· | |
| A | 4278 hCoV-19/Scotland/CVR45/2020\|EPI_ISL_425773\|2020-03 | ···················· | ···················· | ···················· | ···················· | ···················· | ···················· | |
| A | 4279 hCoV-19/Scotland/CVR46/2020\|EPI_ISL_425774\|2020-03 | ···················· | ···················· | ···················· | ···················· | ···················· | ···················· | |
| A | 4280 hCoV-19/Scotland/CVR462/2020\|EPI_ISL_425775\|2020-0 | ···················· | ···················· | ···················· | ···················· | ···················· | ···················· | |
| A | 4281 hCoV-19/Scotland/CVR47/2020\|EPI_ISL_425776\|2020-03 | ···················· | ···················· | ···················· | ···················· | ···················· | ···················· | |
| A | 4282 hCoV-19/Scotland/CVR49/2020\|EPI_ISL_425777\|2020-03 | ···················· | ···················· | ···················· | ···················· | ···················· | ···················· | |
| A | 4283 hCoV-19/Scotland/CVR50/2020\|EPI_ISL_425778\|2020-03 | ···················· | ···················· | ···················· | ···················· | ···················· | ···················· | |
| A | 4284 hCoV-19/Scotland/CVR51/2020\|EPI_ISL_425779\|2020-03 | ···················· | ···················· | ···················· | ···················· | ···················· | ···················· | |
| A | 4285 hCoV-19/Scotland/CVR52/2020\|EPI_ISL_425780\|2020-03 | ···················· | ···················· | ···················· | ···················· | ···················· | ···················· | |
| A | 4286 hCoV-19/Scotland/CVR53/2020\|EPI_ISL_425782\|2020-03 | ···················· | ···················· | ···················· | ···················· | ···················· | ···················· | |
| A | 4287 hCoV-19/Scotland/CVR54/2020\|EPI_ISL_425783\|2020-03 | ···················· | ···················· | ···················· | ···················· | ···················· | ···················· | |
| A | 4288 hCoV-19/Scotland/CVR55/2020\|EPI_ISL_425784\|2020-03 | ···················· | ···················· | ···················· | ···················· | ···················· | ···················· | |
| A | 4289 hCoV-19/Scotland/CVR56/2020\|EPI_ISL_425785\|2020-03 | ···················· | ···················· | ···················· | ···················· | ···················· | ···················· | |
| A | 4290 hCoV-19/Scotland/CVR562/2020\|EPI_ISL_425786\|2020-0 | ···················· | ···················· | ···················· | ···················· | ···················· | ···················· | |
| A | 4291 hCoV-19/Scotland/CVR57/2020\|EPI_ISL_425787\|2020-03 | ···················· | ···················· | ···················· | ···················· | ···················· | ···················· | |
| A | 4292 hCoV-19/Scotland/CVR58/2020\|EPI_ISL_425788\|2020-03 | ···················· | ···················· | ···················· | ···················· | ···················· | ···················· | |
| A | 4293 hCoV-19/Scotland/CVR60/2020\|EPI_ISL_425789\|2020-03 | ···················· | ···················· | ···················· | ···················· | ···················· | ···················· | |
| A | 4294 hCoV-19/Scotland/EDB113/2020\|EPI_ISL_425900\|2020-0 | ···················· | ···················· | ···················· | ···················· | ···················· | ···················· | |
| A | 4296 hCoV-19/Scotland/EDB118/2020\|EPI_ISL_425902\|2020-0 | ···················· | ···················· | ···················· | ···················· | ···················· | ···················· | |
| A | 4298 hCoV-19/Scotland/EDB120/2020\|EPI_ISL_425904\|2020-0 | ···················· | ···················· | ···················· | ···················· | ···················· | ···················· | |
| A | 4300 hCoV-19/Scotland/EDB124/2020\|EPI_ISL_425906\|2020-0 | ···················· | ···················· | ···················· | ···················· | ···················· | ···················· | |
| A | 4301 hCoV-19/Scotland/EDB125/2020\|EPI_ISL_425907\|2020-0 | ···················· | ···················· | ···················· | ···················· | ···················· | ···················· | |
| A | 4303 hCoV-19/Scotland/EDB132/2020\|EPI_ISL_425911\|2020-0 | ···················· | ···················· | ···················· | ···················· | ···················· | ···················· | |
| A | 4304 hCoV-19/Scotland/EDB142/2020\|EPI_ISL_425912\|2020-0 | ···················· | ···················· | ···················· | ···················· | ···················· | ···················· | |
| A | 4306 hCoV-19/Scotland/EDB146/2020\|EPI_ISL_425915\|2020-0 | ···················· | ···················· | ···················· | ···················· | ···················· | ···················· | |
| A | 4307 hCoV-19/Scotland/EDB148/2020\|EPI_ISL_425916\|2020-0 | ···················· | ···················· | ···················· | ···················· | ···················· | ···················· | |
| A | 4308 hCoV-19/Scotland/EDB149/2020\|EPI_ISL_425917\|2020-0 | ···················· | ···················· | ···················· | ···················· | ···················· | ···················· | |
| A | 4309 hCoV-19/Scotland/EDB150/2020\|EPI_ISL_425918\|2020-0 | ···················· | ···················· | ···················· | ···················· | ···················· | ···················· | |
| A | 4310 hCoV-19/Scotland/EDB151/2020\|EPI_ISL_425919\|2020-0 | ···················· | ·A·················· | ···················· | ···················· | ···················· | ···················· | |
| A | 4311 hCoV-19/Scotland/EDB155/2020\|EPI_ISL_425920\|2020-0 | ···················· | ···················· | ···················· | ···················· | ···················· | ···················· | |
| A | 4312 hCoV-19/Scotland/EDB156/2020\|EPI_ISL_425921\|2020-0 | ···················· | ···················· | ···················· | ···················· | ·······N·N····N····· | ·······NN··········· | 1 |
| A | 4316 hCoV-19/Scotland/EDB188/2020\|EPI_ISL_425932\|2020-0 | ···················· | ···················· | ···················· | ···················· | ···················· | ···················· | |
| A | 4317 hCoV-19/Scotland/EDB189/2020\|EPI_ISL_425933\|2020-0 | ···················· | ···················· | ···················· | ···················· | ···················· | ···················· | |
| A | 4320 hCoV-19/Scotland/EDB193/2020\|EPI_ISL_425936\|2020-0 | ···················· | ···················· | ···················· | ···················· | ···················· | ·······NN··········· | |
| A | 4323 hCoV-19/Scotland/EDB198/2020\|EPI_ISL_425939\|2020-0 | ···················· | ···················· | ···················· | ···················· | ···················· | ···················· | |
| A | 4327 hCoV-19/Scotland/EDB212/2020\|EPI_ISL_425943\|2020-0 | ···················· | ···················· | ···················· | ···················· | ···················· | ···················· | |
| A | 4328 hCoV-19/Sweden/RV-FOI-4/2020\|EPI_ISL_428148\|2020-0 | ···················· | ···················· | ···················· | ···················· | ···················· | ···················· | |
| A | 4329 hCoV-19/Denmark/ALAB-SSI361/2020\|EPI_ISL_429471\|20 | ···················· | ···················· | ···················· | ···················· | ···················· | ···················· | |
| A | 4330 hCoV-19/Denmark/ALAB-SSI364/2020\|EPI_ISL_429472\|20 | ···················· | ···················· | ···················· | ···················· | ···················· | ···················· | |
| A | 4331 hCoV-19/Denmark/ALAB-SSI365/2020\|EPI_ISL_429473\|20 | ···················· | ···················· | ···················· | ···················· | ···················· | ···················· | |
| A | 4332 hCoV-19/Denmark/ALAB-SSI366/2020\|EPI_ISL_429474\|20 | ···················· | ···················· | ···················· | ···················· | ···················· | ···················· | |
| A | 4333 hCoV-19/Denmark/ALAB-SSI370/2020\|EPI_ISL_429476\|20 | ···················· | ···················· | ···················· | ···················· | ···················· | ···················· | |
| A | 4334 hCoV-19/Denmark/ALAB-SSI371/2020\|EPI_ISL_429477\|20 | ···················· | ···················· | ···················· | ···················· | ···················· | ···················· | |
| A | 4335 hCoV-19/Denmark/ALAB-SSI372/2020\|EPI_ISL_429478\|20 | ···················· | ···················· | ···················· | ···················· | ···················· | ···················· | |
| A | 4336 hCoV-19/Denmark/ALAB-SSI374/2020\|EPI_ISL_429479\|20 | ···················· | ·A·················· | ···················· | ···················· | ···················· | ···················· | |
| A | 4337 hCoV-19/Denmark/ALAB-SSI375/2020\|EPI_ISL_429480\|20 | ···················· | ···················· | ···················· | ···················· | ···················· | ···················· | |
| A | 4338 hCoV-19/Denmark/ALAB-SSI376/2020\|EPI_ISL_429481\|20 | ···················· | ···················· | ···················· | ···················· | ···················· | ···················· | |
| A | 4339 hCoV-19/Denmark/ALAB-SSI377/2020\|EPI_ISL_429482\|20 | ···················· | ···················· | ···················· | ···················· | ···················· | ···················· | |
| A | 4340 hCoV-19/Denmark/ALAB-SSI383/2020\|EPI_ISL_429483\|20 | ···················· | ···················· | ···················· | ···················· | ···················· | ···················· | |

| | | | | | | | |
|---|---|---|---|---|---|---|---|
| A | 4341 hCoV-19/Denmark/ALAB-SSI384/2020\|EPI_ISL_429484\|20 | .................... | .................... | .................... | .................... | .................... | .................... |
| A | 4342 hCoV-19/Denmark/ALAB-SSI385/2020\|EPI_ISL_429485\|20 | .................... | .................... | .................... | .................... | .................... | .................... |
| A | 4343 hCoV-19/Denmark/ALAB-SSI390/2020\|EPI_ISL_429486\|20 | .................... | .................... | .................... | .................... | .................... | .................... |
| A | 4344 hCoV-19/Denmark/ALAB-SSI393/2020\|EPI_ISL_429488\|20 | .................... | .................... | .................... | .................... | .................... | .................... |
| A | 4345 hCoV-19/Denmark/ALAB-SSI394/2020\|EPI_ISL_429489\|20 | .................... | .................... | .................... | .................... | .................... | .................... |
| A | 4346 hCoV-19/Denmark/ALAB-SSI395/2020\|EPI_ISL_429490\|20 | .................... | .................... | .................... | .................... | .................... | .................... |
| A | 4347 hCoV-19/Denmark/ALAB-SSI399/2020\|EPI_ISL_429492\|20 | .................... | .................... | .................... | .................... | .................... | .................... |
| A | 4348 hCoV-19/Denmark/ALAB-SSI400/2020\|EPI_ISL_429493\|20 | .................... | .................... | .................... | .................... | .................... | .................... |
| A | 4349 hCoV-19/Denmark/ALAB-SSI402/2020\|EPI_ISL_429495\|20 | .................... | .................... | .................... | .................... | .................... | .................... |
| A | 4350 hCoV-19/Denmark/ALAB-SSI403A/2020\|EPI_ISL_429496\|2 | .................... | .................... | .................... | .................... | .................... | .................... |
| A | 4351 hCoV-19/Denmark/ALAB-SSI404/2020\|EPI_ISL_429497\|20 | .................... | .................... | .................... | .................... | .................... | .................... |
| A | 4352 hCoV-19/Denmark/ALAB-SSI405/2020\|EPI_ISL_429499\|20 | .................... | .................... | .................... | .................... | .................... | .................... |
| A | 4353 hCoV-19/Wuhan/WHU01/2020\|EPI_ISL_406716\|2020-01-02 | .................... | .................... | .................... | .................... | .................... | .................... |
| A | 4354 hCoV-19/Wuhan/WHU02/2020\|EPI_ISL_406717\|2020-01-02 | .................... | .................... | .................... | .................... | .................... | .................... |
| A | 4355 hCoV-19/Wuhan/WH01/2019\|EPI_ISL_406798\|2019-12-26 | .................... | .................... | .................... | .................... | .................... | .................... |
| A | 4356 hCoV-19/Guangdong/2020XN4373-P0039/2020\|EPI_ISL_41 | .................... | .................... | .................... | .................... | .................... | .................... |
| A | 4357 hCoV-19/Guangdong/2020XN4433-P0040/2020\|EPI_ISL_41 | .................... | .................... | .................... | .................... | .................... | .................... |
| A | 4358 hCoV-19/Guangdong/2020XN4243-P0035/2020\|EPI_ISL_41 | .................... | .................... | .................... | .................... | .................... | .................... |
| A | 4359 hCoV-19/Guangdong/2020XN4475-P0042/2020\|EPI_ISL_41 | .................... | .................... | .................... | .................... | .................... | .................... |
| A | 4360 hCoV-19/Guangdong/GD2020234-P0023/2020\|EPI_ISL_413 | .................... | .................... | .................... | .................... | .................... | .................... |
| A | 4361 hCoV-19/Guangdong/GD2020233-P0027/2020\|EPI_ISL_413 | .................... | .................... | .................... | .................... | .................... | .................... |
| A | 4362 hCoV-19/Guangdong/2020XN4448-P0002/2020\|EPI_ISL_41 | .................... | .................... | .................... | .................... | .................... | .................... |
| A | 4363 hCoV-19/Guangdong/2020XN4459-P0041/2020\|EPI_ISL_41 | .................... | .................... | .................... | .................... | .................... | .................... |
| A | 4364 hCoV-19/Guangdong/2020XN4273-P0036/2020\|EPI_ISL_41 | .................... | .................... | .................... | .................... | .................... | .................... |
| A | 4365 hCoV-19/Guangdong/GD2020080-P0010/2020\|EPI_ISL_413 | .................... | .................... | .................... | .................... | .................... | .................... |
| A | 4366 hCoV-19/Guangdong/GD2020086-P0021/2020\|EPI_ISL_413 | .................... | .................... | .................... | .................... | .................... | .................... |
| A | 4367 hCoV-19/Guangdong/GD2020087-P0008/2020\|EPI_ISL_413 | .................... | .................... | .................... | .................... | .................... | .................... |
| A | 4368 hCoV-19/Guangdong/GD2020246-P0028/2020\|EPI_ISL_413 | .................... | .................... | .................... | .................... | .................... | .................... |
| A | 4369 hCoV-19/Denmark/SSI-05/2020\|EPI_ISL_416140\|2020-03 | .................... | .................... | .................... | .................... | .................... | .................... |
| A | 4370 hCoV-19/Denmark/SSI-09/2020\|EPI_ISL_416141\|2020-03 | .................... | .................... | .................... | .................... | .................... | .................... |
| A | 4371 hCoV-19/Denmark/SSI-01/2020\|EPI_ISL_416142\|2020-02 | .................... | .................... | .................... | .................... | .................... | .................... |
| A | 4372 hCoV-19/Denmark/SSI-02/2020\|EPI_ISL_416143\|2020-02 | .................... | .................... | .................... | .................... | .................... | .................... |
| A | 4373 hCoV-19/Denmark/SSI-03/2020\|EPI_ISL_416144\|2020-03 | .................... | .................... | .................... | .................... | .................... | .................... |
| A | 4374 hCoV-19/Denmark/SSI-04/2020\|EPI_ISL_416153\|2020-03 | .................... | .................... | .................... | .................... | .................... | .................... |
| A | 4375 hCoV-19/Australia/NSW44/2020\|EPI_ISL_417400\|2020-0 | .................... | .................... | .................... | .................... | .................... | .................... |
| A | 4376 hCoV-19/Australia/NSW45/2020\|EPI_ISL_417401\|2020-0 | .................... | .................... | .................... | .................... | .................... | .................... |
| A | 4377 hCoV-19/Australia/NSW47/2020\|EPI_ISL_417402\|2020-0 | .................... | .................... | .................... | .................... | .................... | .................... |
| A | 4378 hCoV-19/Australia/NSW48/2020\|EPI_ISL_417403\|2020-0 | .................... | .................... | .................... | .................... | .................... | .................... |
| A | 4379 hCoV-19/Australia/NSW50/2020\|EPI_ISL_417404\|2020-0 | .................... | .................... | .................... | .................... | .................... | .................... |
| A | 4380 hCoV-19/Australia/NSW52/2020\|EPI_ISL_417405\|2020-0 | .................... | .................... | .................... | .................... | .................... | .................... |
| A | 4381 hCoV-19/Australia/NSW53/2020\|EPI_ISL_417406\|2020-0 | .................... | .................... | .................... | .................... | .................... | .................... |
| A | 4382 hCoV-19/Australia/NSW54/2020\|EPI_ISL_417407\|2020-0 | .................... | .................... | .................... | .................... | .................... | .................... |
| A | 4383 hCoV-19/Australia/NSW56/2020\|EPI_ISL_417408\|2020-0 | .................... | .................... | .................... | .................... | .................... | .................... |
| A | 4384 hCoV-19/Australia/NSW57/2020\|EPI_ISL_417409\|2020-0 | .................... | .................... | .................... | .................... | .................... | .................... |
| A | 4385 hCoV-19/Australia/NSW58/2020\|EPI_ISL_417410\|2020-0 | .................... | .................... | .................... | .................... | .................... | .................... |
| A | 4386 hCoV-19/Australia/NSW61/2020\|EPI_ISL_417411\|2020-0 | .................... | .................... | .................... | .................... | .................... | .................... |
| A | 4387 hCoV-19/Australia/NSW62/2020\|EPI_ISL_417412\|2020-0 | .................... | .................... | .................... | .................... | .................... | .................... |
| A | 4388 hCoV-19/Italy/FVG-ICGEB-S1/2020\|EPI_ISL_417418\|202 | .................... | .................... | .................... | .................... | .................... | .................... |
| A | 4389 hCoV-19/Italy/FVG-ICGEB-S5/2020\|EPI_ISL_417419\|202 | .................... | .................... | .................... | .................... | .................... | .................... |
| A | 4390 hCoV-19/NanChang/JX216/2020\|EPI_ISL_417420\|2020-03 | .................... | .................... | .................... | .................... | .................... | .................... |
| A | 4391 hCoV-19/Italy/FVG-ICGEB-S8/2020\|EPI_ISL_417421\|202 | .................... | .................... | .................... | .................... | .................... | .................... |
| A | 4392 hCoV-19/Belgium/DHWM-03041/2020\|EPI_ISL_417422\|202 | .................... | .................... | .................... | .................... | .................... | .................... |
| A | 4393 hCoV-19/Belgium/KA-03042/2020\|EPI_ISL_417424\|2020- | .................... | .................... | .................... | .................... | .................... | .................... |
| A | 4394 hCoV-19/Belgium/VBK-03061/2020\|EPI_ISL_417425\|2020 | .................... | .................... | .................... | .................... | .................... | .................... |
| A | 4395 hCoV-19/Belgium/SQ-03043/2020\|EPI_ISL_417426\|2020- | .................... | .................... | .................... | .................... | .................... | .................... |
| A | 4396 hCoV-19/Belgium/VI-03027/2020\|EPI_ISL_417427\|2020- | .................... | .................... | .................... | .................... | .................... | .................... |
| A | 4397 hCoV-19/Belgium/FR-03029/2020\|EPI_ISL_417428\|2020- | .................... | .................... | .................... | .................... | .................... | .................... |
| A | 4398 hCoV-19/Belgium/MMJ-03034/2020\|EPI_ISL_417429\|2020 | .................... | .................... | .................... | .................... | .................... | .................... |
| A | 4399 hCoV-19/Belgium/RT-03035/2020\|EPI_ISL_417430\|2020- | .................... | .................... | .................... | .................... | .................... | .................... |
| A | 4401 hCoV-19/DRC/KN-0038/2020\|EPI_ISL_417434\|2020-03-14 | .................... | .................... | .................... | .................... | .................... | .................... |
| A | 4403 hCoV-19/DRC/KN-0051/2020\|EPI_ISL_417436\|2020-03-17 | .................... | .................... | .................... | .................... | .................... | .................... |
| A | 4404 hCoV-19/DRC/KN-0054/2020\|EPI_ISL_417437\|2020-03-17 | .................... | .................... | .................... | .................... | .................... | .................... |
| A | 4405 hCoV-19/DRC/KN-0058/2020\|EPI_ISL_417438\|2020-03-17 | .................... | .................... | .................... | .................... | .................... | .................... |
| A | 4406 hCoV-19/DRC/KN-0059/2020\|EPI_ISL_417439\|2020-03-17 | .................... | .................... | .................... | .................... | .................... | .................... |
| A | 4407 hCoV-19/DRC/KN-0060/2020\|EPI_ISL_417440\|2020-03-17 | .................... | .................... | .................... | .................... | .................... | .................... |
| A | 4408 hCoV-19/DRC/KN-0070/2020\|EPI_ISL_417441\|2020-03-17 | .................... | .................... | .................... | .................... | .................... | .................... |
| A | 4409 hCoV-19/DRC/KN-0072/2020\|EPI_ISL_417442\|2020-03-18 | .................... | .................... | .................... | .................... | .................... | .................... |
| A | 4410 hCoV-19/Hong Kong/XM-PII-S4/2020\|EPI_ISL_417443\|20 | .................... | .................... | .................... | .................... | .................... | .................... |
| A | 4411 hCoV-19/Pakistan/Gilgit1/2020\|EPI_ISL_417444\|2020- | .................... | .................... | .................... | .................... | .................... | .................... |
| A | 4412 hCoV-19/Italy/LOM-UniMI01/2020\|EPI_ISL_417445\|2020 | .................... | .................... | .................... | .................... | .................... | .................... |
| A | 4413 hCoV-19/Italy/LOM-UniMI03/2020\|EPI_ISL_417447\|2020 | .................... | .................... | .................... | .................... | .................... | .................... |
| A | 4414 hCoV-19/USA/WA-UW234/2020\|EPI_ISL_417448\|2020-03-1 | .................... | .................... | .................... | .................... | .................... | .................... |
| A | 4415 hCoV-19/USA/WA-UW235/2020\|EPI_ISL_417449\|2020-03-1 | .................... | .................... | .................... | .................... | .................... | .................... |
| A | 4416 hCoV-19/USA/WA-UW236/2020\|EPI_ISL_417450\|2020-03-1 | .................... | .................... | .................... | .................... | .................... | .................... |
| A | 4417 hCoV-19/USA/WA-UW237/2020\|EPI_ISL_417451\|2020-03-1 | .................... | .................... | .................... | .................... | .................... | .................... |
| A | 4418 hCoV-19/USA/CT-UW238/2020\|EPI_ISL_417452\|2020-03-1 | .................... | .................... | .................... | .................... | .................... | .................... |
| A | 4419 hCoV-19/USA/WA-UW239/2020\|EPI_ISL_417453\|2020-03-1 | .................... | .................... | .................... | .................... | .................... | .................... |
| A | 4420 hCoV-19/USA/WA-UW240/2020\|EPI_ISL_417454\|2020-03-1 | .................... | .................... | .................... | .................... | .................... | .................... |

A 4421 hCoV-19/USA/WA-UW241/2020|EPI_ISL_417455|2020-03-1 .................... .................... .................... .................... .................... ....................
A 4422 hCoV-19/USA/WA-UW242/2020|EPI_ISL_417456|2020-03-1 .................... .................... .................... .................... .................... ....................
A 4423 hCoV-19/Germany/NW-HHU-12/2020|EPI_ISL_417457|2020 .................... .................... .................... .................... .................... ....................
A 4424 hCoV-19/Germany/NW-HHU-13/2020|EPI_ISL_417458|2020 .................... .................... .................... .................... .................... ....................
A 4425 hCoV-19/Germany/NW-HHU-14/2020|EPI_ISL_417459|2020 .................... .................... .................... .................... .................... ....................
A 4426 hCoV-19/Germany/NW-HHU-15/2020|EPI_ISL_417460|2020 .................... .................... .................... .................... .................... ....................
A 4427 hCoV-19/Germany/NW-HHU-16/2020|EPI_ISL_417461|2020 .................... .................... .................... .................... .................... ....................
A 4428 hCoV-19/Germany/NW-HHU-17/2020|EPI_ISL_417462|2020 .................... .................... .................... .................... .................... ....................
A 4429 hCoV-19/Germany/NW-HHU-18/2020|EPI_ISL_417463|2020 .................... .................... .................... .................... .................... ....................
A 4430 hCoV-19/Germany/NW-HHU-19/2020|EPI_ISL_417464|2020 .................... .................... .................... .................... .................... ....................
A 4431 hCoV-19/Germany/NW-HHU-20/2020|EPI_ISL_417465|2020 .................... .................... .................... .................... .................... ....................
A 4432 hCoV-19/Germany/NW-HHU-21/2020|EPI_ISL_417466|2020 .................... .................... .................... .................... .................... ....................
A 4433 hCoV-19/Germany/NW-HHU-22/2020|EPI_ISL_417467|2020 .................... .................... .................... .................... .................... ....................
A 4434 hCoV-19/Germany/NW-HHU-23/2020|EPI_ISL_417468|2020 .................... .................... .................... .................... .................... ....................
A 4435 hCoV-19/USA/MN31-MDH31/2020|EPI_ISL_417469|2020-03 .................... .................... .................... .................... .................... ....................
A 4436 hCoV-19/USA/MN32-MDH32/2020|EPI_ISL_417470|2020-03 .................... .................... .................... .................... .................... ....................
A 4437 hCoV-19/USA/MN34-MDH34/2020|EPI_ISL_417471|2020-03 .................... .................... .................... .................... .................... ....................
A 4438 hCoV-19/USA/MN35-MDH35/2020|EPI_ISL_417472|2020-03 .................... .................... .................... .................... .................... ....................
A 4439 hCoV-19/USA/MN36-MDH36/2020|EPI_ISL_417473|2020-03 .................... .................... .................... .................... .................... ....................
A 4440 hCoV-19/USA/MN38-MDH38/2020|EPI_ISL_417474|2020-03 .................... .................... .................... .................... .................... ....................
A 4441 hCoV-19/USA/MN39-MDH39/2020|EPI_ISL_417475|2020-03 .................... .................... .................... .................... .................... ....................
A 4442 hCoV-19/USA/MN40-MDH40/2020|EPI_ISL_417476|2020-03 .................... .................... .................... .................... .................... ....................
A 4443 hCoV-19/USA/MN41-MDH41/2020|EPI_ISL_417477|2020-03 .................... .................... .................... .................... .................... ....................
A 4444 hCoV-19/USA/MN42-MDH42/2020|EPI_ISL_417478|2020-03 .................... .................... .................... .................... .................... ....................
A 4445 hCoV-19/USA/MN43-MDH43/2020|EPI_ISL_417479|2020-03 .................... .................... .................... .................... .................... ....................
A 4446 hCoV-19/USA/MN44-MDH44/2020|EPI_ISL_417480|2020-03 .................... .................... .................... .................... .................... ....................
A 4447 hCoV-19/Iceland/LUH-01/2020|EPI_ISL_417481|2020-03 .................... .................... .................... .................... .................... ....................
A 4448 hCoV-19/Norway/1379/2020|EPI_ISL_417483|2020-02-27 .................... .................... .................... .................... .................... ....................
A 4449 hCoV-19/Norway/1380/2020|EPI_ISL_417484|2020-02-26 .................... .................... .................... .................... .................... ....................
A 4450 hCoV-19/Norway/1526/2020|EPI_ISL_417485|2020-03-02 .................... .................... .................... .................... .................... ....................
A 4451 hCoV-19/Norway/1538/2020|EPI_ISL_417486|2020-03-01 .................... .................... .................... .................... .................... ....................
A 4452 hCoV-19/Norway/1539/2020|EPI_ISL_417487|2020-03-01 .................... .................... .................... .................... .................... ....................
A 4453 hCoV-19/Norway/1493/2020|EPI_ISL_417488|2020-02-29 .................... .................... .................... .................... .................... ....................
A 4454 hCoV-19/England/20124013202/2020|EPI_ISL_418700|20 .................... .................... .................... .................... .................... ....................
A 4455 hCoV-19/England/20124020402/2020|EPI_ISL_418701|20 .................... .................... .................... .................... .................... ....................
A 4456 hCoV-19/England/20124020802/2020|EPI_ISL_418702|20 .................... .................... .................... .................... .................... ....................
A 4457 hCoV-19/England/20124021402/2020|EPI_ISL_418704|20 .................... .................... .................... .................... .................... ....................
A 4458 hCoV-19/England/20124029802/2020|EPI_ISL_418706|20 .................... .................... .................... .................... .................... ....................
A 4459 hCoV-19/England/20124030003/2020|EPI_ISL_418707|20 .................... .................... .................... .................... .................... ....................
A 4460 hCoV-19/England/20124030802/2020|EPI_ISL_418708|20 .................... .................... .................... .................... .................... ....................
A 4461 hCoV-19/England/20124034002/2020|EPI_ISL_418709|20 .................... .................... .................... .................... .................... ....................
A 4462 hCoV-19/England/20124034702/2020|EPI_ISL_418711|20 .................... .................... .................... .................... .................... ....................
A 4465 hCoV-19/England/20124036902/2020|EPI_ISL_418718|20 .................... .................... .................... .................... .................... ....................
A 4468 hCoV-19/England/20124049402/2020|EPI_ISL_418723|20 .................... .................... .................... .................... .................... ....................
A 4469 hCoV-19/England/20124095502/2020|EPI_ISL_418729|20 .................... .................... .................... .................... .................... ....................
A 4471 hCoV-19/England/20124097102/2020|EPI_ISL_418734|20 .................... .................... .................... .................... .................... ....................
A 4474 hCoV-19/England/20124101102/2020|EPI_ISL_418739|20 .................... .................... .................... .................... .................... ....................
A 4477 hCoV-19/England/20126007002/2020|EPI_ISL_418750|20 .................... .................... .................... .................... .................... ....................
A 4478 hCoV-19/England/20126007102/2020|EPI_ISL_418751|20 .................... .................... .................... .................... .................... ....................
A 4479 hCoV-19/England/20126012702/2020|EPI_ISL_418756|20 .................... .................... .................... .................... .................... ....................
A 4480 hCoV-19/England/20126035502/2020|EPI_ISL_418764|20 .................... .................... .................... .................... .................... ....................
A 4481 hCoV-19/England/20126096602/2020|EPI_ISL_418770|20 .................... .................... .................... .................... .................... ....................
A 4482 hCoV-19/USA/WA-NH2/2020|EPI_ISL_418771|2020-03-13 .................... .................... .................... .................... .................... ....................
A 4483 hCoV-19/USA/WA-NH3/2020|EPI_ISL_418772|2020-03-13 .................... .................... .................... .................... .................... ....................
A 4484 hCoV-19/USA/WA-NH4/2020|EPI_ISL_418773|2020-03-13 .................... .................... .................... .................... .................... ....................
A 4485 hCoV-19/USA/WA-NH5/2020|EPI_ISL_418774|2020-03-13 .................... .................... .................... .................... .................... ....................
A 4486 hCoV-19/USA/WA-NH6/2020|EPI_ISL_418775|2020-03-13 .................... .................... .................... .................... .................... ....................
A 4487 hCoV-19/USA/WA-NH7/2020|EPI_ISL_418776|2020-03-13 .................... .................... .................... .................... .................... ....................
A 4488 hCoV-19/USA/WA-NH8/2020|EPI_ISL_418777|2020-03-13 .................... .................... .................... .................... .................... ....................
A 4489 hCoV-19/USA/WA-NH9/2020|EPI_ISL_418778|2020-03-13 .................... .................... .................... .................... .................... ....................
A 4490 hCoV-19/USA/WA-NH10/2020|EPI_ISL_418779|2020-03-13 .................... .................... .................... .................... .................... ....................
A 4491 hCoV-19/USA/WA-NH11/2020|EPI_ISL_418780|2020-03-13 .................... .................... .................... .................... .................... ....................
A 4492 hCoV-19/USA/WA-NH12/2020|EPI_ISL_418781|2020-03-13 .................... .................... .................... .................... .................... ....................
A 4493 hCoV-19/USA/WA-NH13/2020|EPI_ISL_418782|2020-03-13 .................... .................... .................... .................... .................... ....................
A 4494 hCoV-19/USA/WA-NH14/2020|EPI_ISL_418783|2020-03-13 .................... .................... .................... .................... .................... ....................
A 4495 hCoV-19/USA/WA-NH17/2020|EPI_ISL_418784|2020-03-13 .................... .................... .................... .................... .................... ....................
A 4496 hCoV-19/USA/WA-NH18/2020|EPI_ISL_418785|2020-03-13 .................... .................... .................... .................... .................... ....................
A 4497 hCoV-19/USA/WA-NH19/2020|EPI_ISL_418786|2020-03-13 .................... .................... .................... .................... .................... ....................
A 4498 hCoV-19/USA/WA-NH20/2020|EPI_ISL_418787|2020-03-13 .................... .................... .................... .................... .................... ....................
A 4499 hCoV-19/USA/WA-NH21/2020|EPI_ISL_418788|2020-03-13 .................... .................... .................... .................... .................... ....................
A 4500 hCoV-19/USA/WA-NH22/2020|EPI_ISL_418789|2020-03-13 .................... .................... .................... .................... .................... ....................
A 4501 hCoV-19/USA/WA-NH23/2020|EPI_ISL_418790|2020-03-13 .................... .................... .................... .................... .................... ....................
A 4502 hCoV-19/USA/WA-NH24/2020|EPI_ISL_418791|2020-03-13 .................... .................... .................... .................... .................... ....................
A 4503 hCoV-19/Belgium/GL-030546/2020|EPI_ISL_418792|2020 .................... .................... .................... .................... .................... ....................
A 4504 hCoV-19/Belgium/SS-031047/2020|EPI_ISL_418793|2020 .................... .................... .................... .................... .................... ....................
A 4505 hCoV-19/Belgium/VHRA-030644/2020|EPI_ISL_418794|20 .................... .................... .................... .................... .................... ....................
A 4506 hCoV-19/Belgium/NKR-030645/2020|EPI_ISL_418795|202 .................... .................... .................... .................... .................... ....................
A 4507 hCoV-19/Belgium/VDLH-030548/2020|EPI_ISL_418796|20 .................... .................... .................... .................... .................... ....................

| | | | | | | | | | |
|---|---|---|---|---|---|---|---|---|---|
| A | 4508 hCoV-19/Belgium/VDLH-030548-2/2020\|EPI_ISL_418797\| | ···················· | ···················· | ···················· | ···················· | ···················· | ···················· | | |
| A | 4509 hCoV-19/Belgium/GS-030549/2020\|EPI_ISL_418798\|2020 | ···················· | ···················· | ···················· | ···················· | ···················· | ···················· | | |
| A | 4510 hCoV-19/Australia/QLDID922/2020\|EPI_ISL_418799\|202 | ···················· | ···················· | ···················· | ···················· | ···················· | ···················· | | |
| A | 4511 hCoV-19/England/201360220/2020\|EPI_ISL_423250\|2020 | ···················· | ···················· | ···················· | ···················· | ···················· | ···················· | | |
| A | 4512 hCoV-19/England/201360221/2020\|EPI_ISL_423251\|2020 | ···················· | ···················· | ···················· | ···················· | ···················· | ···················· | | |
| A | 4513 hCoV-19/England/201360223/2020\|EPI_ISL_423253\|2020 | ···················· | ···················· | ···················· | ···················· | ···················· | ···················· | | |
| A | 4514 hCoV-19/England/201360224/2020\|EPI_ISL_423254\|2020 | ···················· | ···················· | ···················· | ···················· | ···················· | ···················· | | |
| A | 4516 hCoV-19/England/201360236/2020\|EPI_ISL_423256\|2020 | ···················· | ···················· | ···················· | ···················· | ···················· | ···················· | | |
| A | 4518 hCoV-19/England/201360251/2020\|EPI_ISL_423259\|2020 | ···················· | ···················· | ···················· | ···················· | ·N················NN | ··N················· | 1 | 1 |
| A | 4519 hCoV-19/England/201360275/2020\|EPI_ISL_423260\|2020 | ···················· | ···················· | ···················· | ···················· | ···················· | ······R············· | | |
| A | 4520 hCoV-19/England/201360276/2020\|EPI_ISL_423261\|2020 | ···················· | ···················· | ···················· | ···················· | ···················· | ···················· | | |
| A | 4521 hCoV-19/England/201360277/2020\|EPI_ISL_423262\|2020 | ···················· | ···················· | ···················· | ···················· | ···················· | ···················· | | |
| A | 4522 hCoV-19/England/201360281/2020\|EPI_ISL_423263\|2020 | ···················· | ···················· | ···················· | ···················· | ···················· | ···················· | | |
| A | 4523 hCoV-19/England/201360317/2020\|EPI_ISL_423264\|2020 | ···················· | ···················· | ···················· | ···················· | ···················· | ···················· | | |
| A | 4524 hCoV-19/England/201360318/2020\|EPI_ISL_423265\|2020 | ···················· | ···················· | ···················· | ···················· | ···················· | NN·N················ | | 1 |
| A | 4526 hCoV-19/England/20138007104/2020\|EPI_ISL_423269\|20 | ···················· | ···················· | ···················· | ···················· | ···················· | ···N················ | | |
| A | 4528 hCoV-19/England/20138010904/2020\|EPI_ISL_423277\|20 | ···················· | ···················· | ···················· | ···················· | ···················· | ···················· | | |
| A | 4529 hCoV-19/England/20138017804/2020\|EPI_ISL_423288\|20 | ···················· | ···················· | ···················· | ···················· | ···················· | ···················· | | |
| A | 4530 hCoV-19/England/20138019104/2020\|EPI_ISL_423292\|20 | ···················· | ···················· | ···················· | ···················· | ···················· | ···················· | | |
| A | 4531 hCoV-19/Iceland/477/2020\|EPI_ISL_424500\|2020-03-20 | ···················· | ···················· | ···················· | ···················· | ···················· | ···················· | | |
| A | 4532 hCoV-19/Iceland/479/2020\|EPI_ISL_424502\|2020-03-20 | ···················· | ···················· | ···················· | ···················· | ···················· | ···················· | | |
| A | 4533 hCoV-19/Iceland/480/2020\|EPI_ISL_424503\|2020-03-20 | ···················· | ···················· | ···················· | ···················· | ···················· | ···················· | | |
| A | 4534 hCoV-19/Iceland/482/2020\|EPI_ISL_424505\|2020-03-20 | ···················· | ···················· | ···················· | ···················· | ···················· | ···················· | | |
| A | 4535 hCoV-19/Iceland/483/2020\|EPI_ISL_424506\|2020-03-20 | ···················· | ···················· | ···················· | ···················· | ···················· | ···················· | | |
| A | 4536 hCoV-19/Iceland/484/2020\|EPI_ISL_424507\|2020-03-20 | ···················· | ···················· | ···················· | ···················· | ···················· | ···················· | | |
| A | 4537 hCoV-19/Iceland/485/2020\|EPI_ISL_424508\|2020-03-20 | ···················· | ···················· | ···················· | ···················· | ···················· | ···················· | | |
| A | 4538 hCoV-19/Iceland/487/2020\|EPI_ISL_424509\|2020-03-20 | ···················· | ···················· | ···················· | ···················· | ···················· | ···················· | | |
| A | 4539 hCoV-19/Iceland/488/2020\|EPI_ISL_424510\|2020-03-20 | ···················· | ···················· | ···················· | ···················· | ···················· | ···················· | | |
| A | 4540 hCoV-19/Iceland/489/2020\|EPI_ISL_424511\|2020-03-20 | ···················· | ···················· | ···················· | ···················· | ···················· | ···················· | | |
| A | 4541 hCoV-19/Iceland/490/2020\|EPI_ISL_424512\|2020-03-20 | ···················· | ···················· | ···················· | ···················· | ···················· | ···················· | | |
| A | 4542 hCoV-19/Iceland/491/2020\|EPI_ISL_424513\|2020-03-20 | ···················· | ···················· | ···················· | ···················· | ···················· | ···················· | | |
| A | 4543 hCoV-19/Iceland/560/2020\|EPI_ISL_424580\|2020-03-27 | ···················· | ···················· | ···················· | ···················· | ···················· | ···················· | | |
| A | 4544 hCoV-19/Iceland/561/2020\|EPI_ISL_424581\|2020-03-28 | ···················· | ···················· | ···················· | ···················· | ···················· | ···················· | | |
| A | 4545 hCoV-19/Iceland/562/2020\|EPI_ISL_424582\|2020-03-27 | ···················· | ···················· | ···················· | ···················· | ···················· | ···················· | | |
| A | 4546 hCoV-19/Iceland/564/2020\|EPI_ISL_424584\|2020-03-28 | ···················· | ···················· | ···················· | ···················· | ···················· | ···················· | | |
| A | 4547 hCoV-19/Iceland/565/2020\|EPI_ISL_424585\|2020-03-28 | ···················· | ···················· | ···················· | ···················· | ···················· | ···················· | | |
| A | 4548 hCoV-19/Iceland/568/2020\|EPI_ISL_424588\|2020-03-28 | ···················· | ···················· | ···················· | ···················· | ···················· | ···················· | | |
| A | 4549 hCoV-19/Iceland/569/2020\|EPI_ISL_424589\|2020-03-28 | ···················· | ···················· | ···················· | ···················· | ···················· | ···················· | | |
| A | 4550 hCoV-19/Iceland/570/2020\|EPI_ISL_424590\|2020-03-28 | ···················· | ···················· | ···················· | ···················· | ···················· | ···················· | | |
| A | 4551 hCoV-19/Iceland/571/2020\|EPI_ISL_424591\|2020-03-28 | ···················· | ···················· | ···················· | ···················· | ···················· | ···················· | | |
| A | 4552 hCoV-19/Iceland/572/2020\|EPI_ISL_424592\|2020-03-28 | ···················· | ···················· | ···················· | ···················· | ···················· | ···················· | | |
| A | 4553 hCoV-19/Iceland/573/2020\|EPI_ISL_424593\|2020-03-28 | ···················· | ···················· | ···················· | ···················· | ···················· | ···················· | | |
| A | 4554 hCoV-19/Iceland/575/2020\|EPI_ISL_424595\|2020-03-28 | ···················· | ···················· | ···················· | ···················· | ···················· | ···················· | | |
| A | 4555 hCoV-19/Iceland/576/2020\|EPI_ISL_424596\|2020-03-28 | ···················· | ···················· | ···················· | ···················· | ···················· | ···················· | | |
| A | 4556 hCoV-19/Iceland/577/2020\|EPI_ISL_424597\|2020-03-27 | ···················· | ···················· | ···················· | ···················· | ···················· | ···················· | | |
| A | 4557 hCoV-19/Iceland/578/2020\|EPI_ISL_424598\|2020-03-27 | ···················· | ···················· | ···················· | ···················· | ···················· | ···················· | | |
| A | 4558 hCoV-19/Iceland/579/2020\|EPI_ISL_424599\|2020-03-27 | ···················· | ···················· | ···················· | ···················· | ···················· | ···················· | | |
| A | 4559 hCoV-19/Scotland/EDB024/2020\|EPI_ISL_425830\|2020-0 | ···················· | ···················· | ···················· | ···················· | ···················· | NN·················· | | |
| A | 4561 hCoV-19/Scotland/EDB030/2020\|EPI_ISL_425836\|2020-0 | ···················· | ···················· | ···················· | ···················· | ···················· | ···················· | | |
| A | 4562 hCoV-19/Scotland/EDB033/2020\|EPI_ISL_425839\|2020-0 | ···················· | ···················· | ···················· | ···················· | ···················· | ···················· | | |
| A | 4563 hCoV-19/Scotland/EDB036/2020\|EPI_ISL_425842\|2020-0 | ···················· | ···················· | ···················· | ···················· | ···················· | ···················· | | |
| A | 4564 hCoV-19/Scotland/EDB037/2020\|EPI_ISL_425843\|2020-0 | ···················· | ············A······· | ···················· | ···················· | ···················· | ···················· | | |
| A | 4565 hCoV-19/Scotland/EDB038/2020\|EPI_ISL_425844\|2020-0 | ···················· | ···················· | ···················· | ···················· | ···················· | ···················· | | |
| A | 4566 hCoV-19/Scotland/EDB157/2020\|EPI_ISL_425922\|2020-0 | ···················· | ···················· | ···················· | ···················· | ···················· | ···················· | | |
| A | 4568 hCoV-19/Scotland/EDB162/2020\|EPI_ISL_425924\|2020-0 | ···················· | ···················· | ···················· | ···················· | ···················· | ···················· | | |
| A | 4569 hCoV-19/Scotland/EDB168/2020\|EPI_ISL_425925\|2020-0 | ···················· | ···················· | ···················· | ···················· | ···················· | ···················· | | |
| A | 4571 hCoV-19/Scotland/EDB181/2020\|EPI_ISL_425927\|2020-0 | ···················· | ···················· | ···················· | ···················· | ···················· | ···················· | | |
| A | 4573 hCoV-19/Denmark/ALAB-SSI204/2020\|EPI_ISL_429402\|20 | ···················· | ···················· | ···················· | ···················· | ···················· | ···················· | | |
| A | 4574 hCoV-19/Denmark/ALAB-SSI207/2020\|EPI_ISL_429404\|20 | ···················· | ···················· | ···················· | ···················· | ···················· | ···················· | | |
| A | 4575 hCoV-19/Denmark/ALAB-SSI209/2020\|EPI_ISL_429405\|20 | ···················· | ···················· | ···················· | ···················· | ···················· | ···················· | | |
| A | 4576 hCoV-19/Denmark/ALAB-SSI213/2020\|EPI_ISL_429406\|20 | ···················· | ···················· | ···················· | ···················· | ···················· | ···················· | | |
| A | 4577 hCoV-19/Denmark/ALAB-SSI215/2020\|EPI_ISL_429407\|20 | ···················· | ···················· | ···················· | ···················· | ···················· | ···················· | | |
| A | 4578 hCoV-19/Denmark/ALAB-SSI216/2020\|EPI_ISL_429408\|20 | ···················· | ···················· | ···················· | ···················· | ···················· | ···················· | | |
| A | 4579 hCoV-19/Denmark/ALAB-SSI219/2020\|EPI_ISL_429409\|20 | ···················· | ···················· | ···················· | ···················· | ···················· | ···················· | | |
| A | 4580 hCoV-19/Denmark/ALAB-SSI221/2020\|EPI_ISL_429410\|20 | ···················· | ···················· | ···················· | ···················· | ···················· | ···················· | | |
| A | 4581 hCoV-19/Denmark/ALAB-SSI235/2020\|EPI_ISL_429414\|20 | ···················· | ···················· | ···················· | ···················· | ···················· | ···················· | | |
| A | 4582 hCoV-19/Denmark/ALAB-SSI258/2020\|EPI_ISL_429420\|20 | ···················· | ···················· | ···················· | ···················· | ···················· | ···················· | | |
| A | 4583 hCoV-19/Denmark/ALAB-SSI259/2020\|EPI_ISL_429421\|20 | ···················· | ···················· | ···················· | ···················· | ···················· | ···················· | | |
| A | 4584 hCoV-19/Denmark/ALAB-SSI262/2020\|EPI_ISL_429424\|20 | ···················· | ···················· | ···················· | ···················· | ···················· | ···················· | | |
| A | 4585 hCoV-19/Denmark/ALAB-SSI263/2020\|EPI_ISL_429425\|20 | ···················· | ···················· | ···················· | ···················· | ···················· | ···················· | | |
| A | 4586 hCoV-19/Denmark/ALAB-SSI264/2020\|EPI_ISL_429426\|20 | ···················· | ···················· | ···················· | ···················· | ···················· | ···················· | | |
| A | 4587 hCoV-19/Denmark/ALAB-SSI265/2020\|EPI_ISL_429427\|20 | ···················· | ···················· | ···················· | ···················· | ···················· | ···················· | | |
| A | 4588 hCoV-19/Denmark/ALAB-SSI268/2020\|EPI_ISL_429428\|20 | ···················· | ···················· | ···················· | ···················· | ···················· | ···················· | | |
| A | 4589 hCoV-19/Denmark/ALAB-SSI271/2020\|EPI_ISL_429431\|20 | ···················· | ···················· | ···················· | ···················· | ···················· | ···················· | | |
| A | 4590 hCoV-19/Denmark/ALAB-SSI272/2020\|EPI_ISL_429433\|20 | ···················· | ···················· | ···················· | ···················· | ···················· | ···················· | | |
| A | 4591 hCoV-19/Denmark/ALAB-SSI273/2020\|EPI_ISL_429434\|20 | ···················· | ···················· | ···················· | ···················· | ···················· | ···················· | | |
| A | 4592 hCoV-19/Denmark/ALAB-SSI279/2020\|EPI_ISL_429439\|20 | ···················· | ···················· | ···················· | ···················· | ···················· | ···················· | | |
| A | 4593 hCoV-19/Denmark/ALAB-SSI280/2020\|EPI_ISL_429440\|20 | ···················· | ···················· | ···················· | ···················· | ···················· | ···················· | | |

A 4594 hCoV-19/Denmark/ALAB-SSI29/2020|EPI_ISL_429447|202 .................... .................... .................... .................... .................... ....................
A 4595 hCoV-19/Denmark/ALAB-SSI293/2020|EPI_ISL_429448|20 .................... .................... .................... .................... .................... ....................
A 4596 hCoV-19/Denmark/ALAB-SSI298/2020|EPI_ISL_429449|20 .................... .................... .................... .................... .................... ....................
A 4597 hCoV-19/Denmark/ALAB-SSI299/2020|EPI_ISL_429450|20 .................... .................... .................... .................... .................... ....................
A 4598 hCoV-19/Denmark/ALAB-SSI300/2020|EPI_ISL_429451|20 .................... .................... .................... .................... .................... ....................
A 4599 hCoV-19/Denmark/ALAB-SSI301/2020|EPI_ISL_429452|20 .................... .................... .................... .................... .................... ....................
A 4600 hCoV-19/Denmark/ALAB-SSI302/2020|EPI_ISL_429453|20 .................... .................... .................... .................... .................... ....................
A 4601 hCoV-19/Denmark/ALAB-SSI346/2020|EPI_ISL_429460|20 .................... .................... .................... .................... .................... ....................
A 4602 hCoV-19/Denmark/ALAB-SSI347/2020|EPI_ISL_429461|20 .................... .................... .................... .................... .................... ....................
A 4603 hCoV-19/Denmark/ALAB-SSI349/2020|EPI_ISL_429463|20 .................... .................... .................... .................... .................... ....................
A 4604 hCoV-19/Denmark/ALAB-SSI351/2020|EPI_ISL_429464|20 .................... .................... .................... .................... .................... ....................
A 4605 hCoV-19/Denmark/ALAB-SSI352/2020|EPI_ISL_429465|20 .................... .................... .................... .................... .................... ....................
A 4606 hCoV-19/Denmark/ALAB-SSI354/2020|EPI_ISL_429466|20 .................... .................... .................... .................... .................... ....................
A 4607 hCoV-19/Denmark/ALAB-SSI357/2020|EPI_ISL_429468|20 .................... .................... .................... .................... .................... ....................
A 4608 hCoV-19/Denmark/ALAB-SSI358/2020|EPI_ISL_429469|20 .................... .................... .................... .................... .................... ....................
A 4609 hCoV-19/Japan/NA-20-05-1/2020|EPI_ISL_410531|2020- .................... .................... .................... .................... .................... ....................
A 4610 hCoV-19/Japan/OS-20-07-1/2020|EPI_ISL_410532|2020- .................... .................... .................... .................... .................... ....................
A 4611 hCoV-19/Singapore/4/2020|EPI_ISL_410535|2020-02-03 .................... .................... .................... .................... .................... ....................
A 4612 hCoV-19/Singapore/5/2020|EPI_ISL_410536|2020-02-06 .................... .................... .................... .................... .................... ....................
A 4613 hCoV-19/Singapore/6/2020|EPI_ISL_410537|2020-02-09 .................... .................... .................... .................... .................... ....................
A 4614 hCoV-19/Italy/LAZ-INMI1-isl/2020|EPI_ISL_410545|20 .................... .................... .................... .................... .................... ....................
A 4615 hCoV-19/Italy/LAZ-INMI1-cs/2020|EPI_ISL_410546|202 .................... .................... .................... .................... .................... ....................
A 4616 hCoV-19/France/IDF-0571/2020|EPI_ISL_411218|2020-0 .................... .................... .................... .................... .................... ....................
A 4617 hCoV-19/France/IDF-0386-islP1/2020|EPI_ISL_411219| .................... .................... .................... .................... .................... ....................
A 4618 hCoV-19/France/IDF-0386-islP3/2020|EPI_ISL_411220| .................... .................... .................... .................... .................... ....................
A 4619 hCoV-19/Guangdong/GDSZ202008-P0020/2020|EPI_ISL_41 .................... .................... .................... .................... .................... ....................
A 4620 hCoV-19/Guangdong/GDSZ202015-P0019/2020|EPI_ISL_41 .................... .................... .................... .................... .................... ....................
A 4621 hCoV-19/Guangdong/GDFS2020052-P0025/2020|EPI_ISL_4 .................... .................... .................... .................... .................... ....................
A 4622 hCoV-19/Guangdong/GD2020115-P0009/2020|EPI_ISL_413 .................... .................... .................... .................... .................... ....................
A 4623 hCoV-19/Belgium/ULG-10026/2020|EPI_ISL_421200|2020 .................... .................... .................... .................... .................... ....................
A 4624 hCoV-19/Belgium/ULG-10027/2020|EPI_ISL_421201|2020 .................... .................... .................... .................... .................... ....................
A 4625 hCoV-19/Belgium/ULG-10028/2020|EPI_ISL_421202|2020 .................... .................... .................... .................... .................... ....................
A 4626 hCoV-19/Hangzhou/HZCDC6789/2020|EPI_ISL_421221|202 .................... .................... .................... .................... .................... ....................
A 4627 hCoV-19/Hangzhou/HZCDC0162/2020|EPI_ISL_421224|202 .................... .................... .................... .................... .................... ....................
A 4628 hCoV-19/USA/NY-PV09126/2020|EPI_ISL_422500|2020-03 .................... .................... .................... .................... .................... ....................
A 4629 hCoV-19/USA/NY-PV09127/2020|EPI_ISL_422501|2020-03 .................... .................... .................... .................... .................... ....................
A 4630 hCoV-19/USA/NY-PV09128/2020|EPI_ISL_422502|2020-03 .................... .................... .................... .................... .................... ....................
A 4631 hCoV-19/USA/NY-PV09129/2020|EPI_ISL_422503|2020-03 .................... .................... .................... .................... .................... ....................
A 4632 hCoV-19/USA/NY-PV09130/2020|EPI_ISL_422504|2020-03 .................... .................... .................... .................... .................... ....................
A 4633 hCoV-19/USA/NY-PV09132/2020|EPI_ISL_422505|2020-03 .................... .................... .................... .................... .................... ....................
A 4634 hCoV-19/USA/NY-PV09134/2020|EPI_ISL_422506|2020-03 .................... .................... .................... .................... .................... ....................
A 4635 hCoV-19/USA/NY-PV09135/2020|EPI_ISL_422507|2020-03 .................... .................... .................... .................... .................... ....................
A 4636 hCoV-19/USA/NY-PV09137/2020|EPI_ISL_422508|2020-03 .................... .................... .................... .................... .................... ....................
A 4637 hCoV-19/USA/NY-PV09138/2020|EPI_ISL_422509|2020-03 .................... .................... .................... .................... .................... ....................
A 4638 hCoV-19/USA/NY-PV09139/2020|EPI_ISL_422510|2020-03 .................... .................... .................... .................... .................... ....................
A 4639 hCoV-19/USA/NY-PV09140/2020|EPI_ISL_422511|2020-03 .................... .................... .................... .................... .................... ....................
A 4640 hCoV-19/USA/NY-PV09142/2020|EPI_ISL_422513|2020-03 .................... .................... .................... .................... .................... ....................
A 4641 hCoV-19/USA/NY-PV09143/2020|EPI_ISL_422514|2020-03 .................... .................... .................... .................... .................... ....................
A 4642 hCoV-19/USA/NY-PV09144/2020|EPI_ISL_422515|2020-03 .................... .................... .................... .................... .................... ....................
A 4643 hCoV-19/USA/NY-PV09145/2020|EPI_ISL_422516|2020-03 .................... .................... .................... .................... .................... ....................
A 4644 hCoV-19/USA/NY-PV09147/2020|EPI_ISL_422517|2020-03 .................... .................... .................... .................... .................... ....................
A 4645 hCoV-19/USA/NY-PV09151/2020|EPI_ISL_422519|2020-03 .................... .................... .................... .................... .................... ....................
A 4646 hCoV-19/USA/NY-PV09152/2020|EPI_ISL_422520|2020-03 .................... .................... .................... .................... .................... ....................
A 4647 hCoV-19/USA/NY-PV09153/2020|EPI_ISL_422521|2020-03 .................... .................... .................... .................... .................... ....................
A 4648 hCoV-19/USA/NY-PV09154/2020|EPI_ISL_422522|2020-03 .................... .................... .................... .................... .................... ....................
A 4649 hCoV-19/USA/NY-PV09156/2020|EPI_ISL_422523|2020-03 .................... .................... .................... .................... .................... ....................
A 4650 hCoV-19/USA/NY-PV09157/2020|EPI_ISL_422524|2020-03 .................... .................... .................... .................... .................... ....................
A 4651 hCoV-19/USA/NY-PV09158/2020|EPI_ISL_422525|2020-03 .................... .................... .................... .................... .................... ....................
A 4652 hCoV-19/USA/NY-PV09163/2020|EPI_ISL_422527|2020-03 .................... .................... .................... .................... .................... ....................
A 4653 hCoV-19/USA/NY-PV09164/2020|EPI_ISL_422528|2020-03 .................... .................... .................... .................... .................... ....................
A 4654 hCoV-19/USA/NY-PV09165/2020|EPI_ISL_422529|2020-03 .................... .................... .................... .................... .................... ....................
A 4655 hCoV-19/USA/NY-PV09168/2020|EPI_ISL_422530|2020-03 .................... .................... .................... .................... .................... ....................
A 4656 hCoV-19/USA/NY-PV09169/2020|EPI_ISL_422531|2020-03 .................... .................... .................... .................... .................... ....................
A 4657 hCoV-19/USA/NY-PV09170/2020|EPI_ISL_422532|2020-03 .................... .................... .................... .................... .................... ....................
A 4658 hCoV-19/USA/NY-PV09171/2020|EPI_ISL_422533|2020-03 .................... .................... .................... .................... .................... ....................
A 4659 hCoV-19/USA/NY-PV09197/2020|EPI_ISL_422552|2020-03 .................... .................... .................... .................... .................... ....................
A 4660 hCoV-19/USA/NY-PV09198/2020|EPI_ISL_422553|2020-03 .................... .................... .................... .................... .................... ....................
A 4661 hCoV-19/USA/NY-PV09199/2020|EPI_ISL_422554|2020-03 .................... .................... .................... .................... .................... ....................
A 4662 hCoV-19/USA/NY-PV09200/2020|EPI_ISL_422555|2020-03 .................... .................... .................... .................... .................... ....................
A 4663 hCoV-19/England/201360051/2020|EPI_ISL_423201|2020 .................... .................... .................... .................... .................... ....................
A 4665 hCoV-19/England/201360053/2020|EPI_ISL_423203|2020 .................... .................... .................... .................... .................... ....................
A 4666 hCoV-19/England/201360055/2020|EPI_ISL_423204|2020 .................... .................... .................... .................... .................... ....................
A 4670 hCoV-19/England/201360075/2020|EPI_ISL_423210|2020 .................... .................... .................... .................... .................... ....................
A 4671 hCoV-19/England/201360119/2020|EPI_ISL_423214|2020 .................... .................... .................... .................... .................... ....................
A 4672 hCoV-19/England/20136012004/2020|EPI_ISL_423215|20 .................... .................... .................... .................... .................... ....................
A 4673 hCoV-19/England/201360121/2020|EPI_ISL_423216|2020 .................... .................... .................... .................... .................... ....................
A 4674 hCoV-19/England/201360126/2020|EPI_ISL_423218|2020 .................... .................... .................... .................... .................... ....................
A 4675 hCoV-19/England/201360127/2020|EPI_ISL_423219|2020 .................... .................... .................... .................... .................... ....................

| | | | | | | | | | | |
|---|---|---|---|---|---|---|---|---|---|---|
| A | 4676 | hCoV-19/England/201360131/2020\|EPI_ISL_423221\|2020 | ···················· | ···················· | ···················· | ···················· | ···················· | ···················· | | |
| A | 4677 | hCoV-19/England/201360136/2020\|EPI_ISL_423223\|2020 | ···················· | ···················· | ···················· | ···················· | ··N················· | ···················· | | |
| A | 4678 | hCoV-19/England/201360137/2020\|EPI_ISL_423224\|2020 | ···················· | ···················· | ···················· | ···················· | ···················· | ···················· | | |
| A | 4679 | hCoV-19/England/201360143/2020\|EPI_ISL_423227\|2020 | ···················· | ···················· | ···················· | ···················· | ···················· | ···················· | | |
| A | 4680 | hCoV-19/England/201360171/2020\|EPI_ISL_423228\|2020 | ···················· | ···················· | ···················· | ···················· | ···················· | ···················· | | |
| A | 4681 | hCoV-19/England/201360178/2020\|EPI_ISL_423231\|2020 | ···················· | ···················· | ···················· | ···················· | ···················· | ···················· | | |
| A | 4682 | hCoV-19/England/201360181/2020\|EPI_ISL_423232\|2020 | ···················· | ···················· | ···················· | ···················· | ···················· | ···················· | | |
| A | 4684 | hCoV-19/England/201360191/2020\|EPI_ISL_423235\|2020 | ···················· | ···················· | ···················· | ···················· | ···················· | ···················· | | |
| A | 4687 | hCoV-19/England/201360201/2020\|EPI_ISL_423238\|2020 | ···················· | ···················· | ···················· | ···················· | ···················· | ···················· | | |
| A | 4689 | hCoV-19/England/201360203/2020\|EPI_ISL_423240\|2020 | ···················· | ···················· | ···················· | ···················· | ···················· | ···················· | | |
| A | 4690 | hCoV-19/England/201360206/2020\|EPI_ISL_423241\|2020 | ···················· | ···················· | ···················· | ···················· | ···················· | ·NN················· | | |
| A | 4691 | hCoV-19/England/201360210/2020\|EPI_ISL_423242\|2020 | ···················· | ···················· | ···················· | ···················· | ···················· | ···················· | | |
| A | 4692 | hCoV-19/England/201360211/2020\|EPI_ISL_423243\|2020 | ···················· | ···················· | ···················· | ···················· | ···················· | ···················· | | |
| A | 4693 | hCoV-19/England/201360213/2020\|EPI_ISL_423244\|2020 | ···················· | ···················· | ···················· | ···················· | ···················· | ···················· | | |
| A | 4694 | hCoV-19/England/201360215/2020\|EPI_ISL_423246\|2020 | ···················· | ···················· | ···················· | ···················· | ···················· | ···················· | | |
| A | 4696 | hCoV-19/England/20132080004/2020\|EPI_ISL_423803\|20 | ···················· | ···················· | ···················· | ···················· | ···················· | ···················· | | |
| A | 4698 | hCoV-19/England/20132080804/2020\|EPI_ISL_423808\|20 | ···················· | ···················· | ···················· | ···················· | ···················· | ···················· | | |
| A | 4699 | hCoV-19/England/20132081104/2020\|EPI_ISL_423809\|20 | ···················· | ···················· | ···················· | ···················· | ···················· | ···················· | | |
| A | 4703 | hCoV-19/England/20132082004/2020\|EPI_ISL_423813\|20 | ···················· | ···················· | ···················· | ···················· | ···················· | ···················· | | |
| A | 4704 | hCoV-19/England/20132082504/2020\|EPI_ISL_423817\|20 | ···················· | ···················· | ···················· | ···················· | ···················· | ···················· | | |
| A | 4705 | hCoV-19/England/20132090404/2020\|EPI_ISL_423818\|20 | ···················· | ···················· | ···················· | ···················· | ···················· | ···················· | | |
| A | 4706 | hCoV-19/England/20132090504/2020\|EPI_ISL_423819\|20 | ···················· | ···················· | ···················· | ···················· | ···················· | ···················· | | |
| A | 4708 | hCoV-19/England/20132091704/2020\|EPI_ISL_423823\|20 | ···················· | ···················· | ···················· | ···················· | ···················· | ···················· | | |
| A | 4709 | hCoV-19/England/20132091804/2020\|EPI_ISL_423824\|20 | ···················· | ···················· | ···················· | ···················· | ···················· | ···················· | | |
| A | 4710 | hCoV-19/England/20132091904/2020\|EPI_ISL_423825\|20 | ···················· | ···················· | ···················· | ···················· | ···················· | ·N·········N·N······ | | 1 |
| A | 4711 | hCoV-19/England/20132095704/2020\|EPI_ISL_423828\|20 | ···················· | ···················· | ···················· | ···················· | ···················· | ···················· | | |
| A | 4712 | hCoV-19/England/20132096004/2020\|EPI_ISL_423829\|20 | ···················· | ···················· | ···················· | ···················· | ···················· | ···················· | | |
| A | 4713 | hCoV-19/England/20132096304/2020\|EPI_ISL_423830\|20 | ···················· | ···················· | ···················· | ···················· | ···················· | ···················· | | |
| A | 4715 | hCoV-19/England/20132100404/2020\|EPI_ISL_423834\|20 | ···················· | ···················· | ···················· | ···················· | ···················· | ···················· | | |
| A | 4716 | hCoV-19/England/20132100704/2020\|EPI_ISL_423835\|20 | ···················· | ···················· | ···················· | ···················· | ···················· | ···················· | | |
| A | 4717 | hCoV-19/England/20132104104/2020\|EPI_ISL_423837\|20 | ···················· | ···················· | ···················· | ···················· | ···················· | ···················· | | |
| A | 4718 | hCoV-19/England/20132104404/2020\|EPI_ISL_423838\|20 | ···················· | ···················· | ···················· | ···················· | ···················· | ···················· | | |
| A | 4719 | hCoV-19/England/20132105704/2020\|EPI_ISL_423839\|20 | ···················· | ···················· | ···················· | ···················· | ···················· | ···················· | | |
| A | 4720 | hCoV-19/England/20132106104/2020\|EPI_ISL_423841\|20 | ···················· | ···················· | ···················· | ···················· | ···················· | ···················· | | |
| A | 4721 | hCoV-19/England/20132107304/2020\|EPI_ISL_423844\|20 | ···················· | ···················· | ···················· | ···················· | ···················· | ···················· | | |
| A | 4722 | hCoV-19/England/20132107704/2020\|EPI_ISL_423845\|20 | ···················· | ···················· | ···················· | ···················· | ···················· | ···················· | | |
| A | 4723 | hCoV-19/England/20132108004/2020\|EPI_ISL_423847\|20 | ···················· | ···················· | ···················· | ···················· | ···················· | ···················· | | |
| A | 4725 | hCoV-19/England/20132108704/2020\|EPI_ISL_423852\|20 | ···················· | ···················· | ···················· | ···················· | ···················· | ···················· | | |
| A | 4726 | hCoV-19/England/20124036306/2020\|EPI_ISL_423857\|20 | ···················· | ···················· | ···················· | ···················· | ···················· | ···················· | | |
| A | 4727 | hCoV-19/England/20126000104/2020\|EPI_ISL_423859\|20 | ···················· | ···················· | ···················· | ···················· | ···················· | ···················· | | |
| A | 4728 | hCoV-19/England/20126000404/2020\|EPI_ISL_423861\|20 | ···················· | ···················· | ···················· | ···················· | ···················· | ···················· | | |
| A | 4730 | hCoV-19/England/20126001204/2020\|EPI_ISL_423863\|20 | ···················· | ···················· | ···················· | ···················· | ···················· | ··NN················ | | |
| A | 4731 | hCoV-19/England/20126001404/2020\|EPI_ISL_423864\|20 | ···················· | ···················· | ···················· | ···················· | ·················NNN | ···················· | 1 | 1 |
| A | 4732 | hCoV-19/England/20126009204/2020\|EPI_ISL_423883\|20 | ···················· | ···················· | ···················· | ···················· | ···················· | ···················· | | |
| A | 4733 | hCoV-19/Iceland/492/2020\|EPI_ISL_424514\|2020-03-20 | ···················· | ···················· | ···················· | ···················· | ···················· | ···················· | | |
| A | 4734 | hCoV-19/Iceland/493/2020\|EPI_ISL_424515\|2020-03-20 | ···················· | ···················· | ···················· | ···················· | ···················· | ···················· | | |
| A | 4735 | hCoV-19/Iceland/494/2020\|EPI_ISL_424516\|2020-03-20 | ···················· | ···················· | ···················· | ···················· | ···················· | ···················· | | |
| A | 4736 | hCoV-19/Iceland/495/2020\|EPI_ISL_424517\|2020-03-21 | ···················· | ···················· | ···················· | ···················· | ···················· | ···················· | | |
| A | 4737 | hCoV-19/Iceland/496/2020\|EPI_ISL_424518\|2020-03-21 | ···················· | ···················· | ···················· | ···················· | ···················· | ···················· | | |
| A | 4738 | hCoV-19/Iceland/497/2020\|EPI_ISL_424519\|2020-03-21 | ···················· | ···················· | ···················· | ···················· | ···················· | ···················· | | |
| A | 4739 | hCoV-19/Iceland/498/2020\|EPI_ISL_424520\|2020-03-21 | ···················· | ···················· | ···················· | ···················· | ···················· | ···················· | | |
| A | 4740 | hCoV-19/Iceland/499/2020\|EPI_ISL_424521\|2020-03-21 | ···················· | ···················· | ···················· | ···················· | ···················· | ···················· | | |
| A | 4741 | hCoV-19/Iceland/500/2020\|EPI_ISL_424522\|2020-03-21 | ···················· | ···················· | ···················· | ···················· | ···················· | ···················· | | |
| A | 4742 | hCoV-19/Iceland/501/2020\|EPI_ISL_424523\|2020-03-21 | ···················· | ···················· | ···················· | ···················· | ···················· | ···················· | | |
| A | 4743 | hCoV-19/Iceland/502/2020\|EPI_ISL_424524\|2020-03-21 | ···················· | ···················· | ···················· | ···················· | ···················· | ···················· | | |
| A | 4744 | hCoV-19/Iceland/503/2020\|EPI_ISL_424525\|2020-03-21 | ···················· | ···················· | ···················· | ···················· | ···················· | ···················· | | |
| A | 4745 | hCoV-19/Iceland/504/2020\|EPI_ISL_424526\|2020-03-21 | ···················· | ···················· | ···················· | ···················· | ···················· | ···················· | | |
| A | 4746 | hCoV-19/Iceland/505/2020\|EPI_ISL_424527\|2020-03-21 | ···················· | ···················· | ···················· | ···················· | ···················· | ···················· | | |
| A | 4747 | hCoV-19/Iceland/506/2020\|EPI_ISL_424528\|2020-03-21 | ···················· | ···················· | ···················· | ···················· | ···················· | ···················· | | |
| A | 4748 | hCoV-19/Iceland/507/2020\|EPI_ISL_424529\|2020-03-20 | ···················· | ···················· | ···················· | ···················· | ···················· | ···················· | | |
| A | 4749 | hCoV-19/Iceland/508/2020\|EPI_ISL_424530\|2020-03-20 | ···················· | ···················· | ···················· | ···················· | ···················· | ···················· | | |
| A | 4750 | hCoV-19/Iceland/509/2020\|EPI_ISL_424531\|2020-03-20 | ···················· | ···················· | ···················· | ···················· | ···················· | ···················· | | |
| A | 4751 | hCoV-19/Iceland/510/2020\|EPI_ISL_424532\|2020-03-20 | ···················· | ···················· | ···················· | ···················· | ···················· | ···················· | | |
| A | 4752 | hCoV-19/Iceland/511/2020\|EPI_ISL_424533\|2020-03-20 | ···················· | ···················· | ···················· | ···················· | ···················· | ···················· | | |
| A | 4753 | hCoV-19/Iceland/512/2020\|EPI_ISL_424534\|2020-03 | ···················· | ···················· | ···················· | ···················· | ···················· | ···················· | | |
| A | 4754 | hCoV-19/Iceland/513/2020\|EPI_ISL_424535\|2020-03-21 | ···················· | ···················· | ···················· | ···················· | ···················· | ···················· | | |
| A | 4755 | hCoV-19/Iceland/514/2020\|EPI_ISL_424536\|2020-03-21 | ···················· | ···················· | ···················· | ···················· | ···················· | ···················· | | |
| A | 4756 | hCoV-19/Iceland/515/2020\|EPI_ISL_424537\|2020-03 | ···················· | ···················· | ···················· | ···················· | ···················· | ···················· | | |
| A | 4757 | hCoV-19/Iceland/516/2020\|EPI_ISL_424538\|2020-03-24 | ···················· | ···················· | ···················· | ···················· | ···················· | ···················· | | |
| A | 4758 | hCoV-19/Iceland/519/2020\|EPI_ISL_424540\|2020-03 | ···················· | ···················· | ···················· | ···················· | ···················· | ···················· | | |
| A | 4759 | hCoV-19/Iceland/520/2020\|EPI_ISL_424541\|2020-03 | ···················· | ···················· | ···················· | ···················· | ···················· | ···················· | | |
| A | 4760 | hCoV-19/Iceland/521/2020\|EPI_ISL_424542\|2020-03 | ···················· | ···················· | ···················· | ···················· | ···················· | ···················· | | |
| A | 4761 | hCoV-19/Iceland/522/2020\|EPI_ISL_424543\|2020-03-25 | ···················· | ···················· | ···················· | ···················· | ···················· | ···················· | | |
| A | 4762 | hCoV-19/Iceland/524/2020\|EPI_ISL_424545\|2020-03-25 | ···················· | ···················· | ···················· | ···················· | ···················· | ···················· | | |
| A | 4763 | hCoV-19/Iceland/525/2020\|EPI_ISL_424546\|2020-03-25 | ···················· | ···················· | ···················· | ···················· | ···················· | ···················· | | |
| A | 4764 | hCoV-19/Iceland/526/2020\|EPI_ISL_424547\|2020-03 | ···················· | ···················· | ···················· | ···················· | ···················· | ···················· | | |
| A | 4765 | hCoV-19/Iceland/527/2020\|EPI_ISL_424548\|2020-03 | ···················· | ···················· | ···················· | ···················· | ···················· | ···················· | | |
| A | 4766 | hCoV-19/Iceland/528/2020\|EPI_ISL_424549\|2020-03 | ···················· | ···················· | ···················· | ···················· | ···················· | ···················· | | |

| | | | | | | | | |
|---|---|---|---|---|---|---|---|---|
| A | 4767 hCoV-19/Iceland/529/2020\|EPI_ISL_424550\|2020-03 | ···················· | ···················· | ···················· | ···················· | ···················· | ···················· | |
| A | 4768 hCoV-19/Iceland/530/2020\|EPI_ISL_424551\|2020-03-25 | ···················· | ···················· | ···················· | ···················· | ···················· | ···················· | |
| A | 4769 hCoV-19/Iceland/535/2020\|EPI_ISL_424555\|2020-03-29 | ···················· | ···················· | ···················· | ···················· | ···················· | ···················· | |
| A | 4770 hCoV-19/Iceland/536/2020\|EPI_ISL_424556\|2020-03-29 | ···················· | ···················· | ···················· | ···················· | ···················· | ···················· | |
| A | 4771 hCoV-19/Iceland/537/2020\|EPI_ISL_424557\|2020-03-27 | ···················· | ···················· | ···················· | ···················· | ···················· | ···················· | |
| A | 4772 hCoV-19/Iceland/538/2020\|EPI_ISL_424558\|2020-03-27 | ···················· | ···················· | ···················· | ···················· | ···················· | ···················· | |
| A | 4773 hCoV-19/Iceland/539/2020\|EPI_ISL_424559\|2020-03-27 | ···················· | ···················· | ···················· | ···················· | ···················· | ···················· | |
| A | 4774 hCoV-19/Iceland/540/2020\|EPI_ISL_424560\|2020-03-27 | ···················· | ···················· | ···················· | ···················· | ···················· | ···················· | |
| A | 4775 hCoV-19/Iceland/542/2020\|EPI_ISL_424562\|2020-03-26 | ···················· | ···················· | ···················· | ···················· | ···················· | ···················· | |
| A | 4776 hCoV-19/Iceland/543/2020\|EPI_ISL_424563\|2020-03-27 | ···················· | ···················· | ···················· | ···················· | ···················· | ···················· | |
| A | 4777 hCoV-19/Iceland/544/2020\|EPI_ISL_424564\|2020-03-27 | ···················· | ···················· | ···················· | ···················· | ···················· | ···················· | |
| A | 4778 hCoV-19/Iceland/547/2020\|EPI_ISL_424567\|2020-03-28 | ···················· | ···················· | ···················· | ···················· | ···················· | ···················· | |
| A | 4779 hCoV-19/Iceland/548/2020\|EPI_ISL_424568\|2020-03-28 | ···················· | ···················· | ···················· | ···················· | ···················· | ···················· | |
| A | 4780 hCoV-19/Iceland/550/2020\|EPI_ISL_424570\|2020-03-28 | ···················· | ···················· | ···················· | ···················· | ···················· | ···················· | |
| A | 4781 hCoV-19/Iceland/551/2020\|EPI_ISL_424571\|2020-03-27 | ···················· | ···················· | ···················· | ···················· | ···················· | ···················· | |
| A | 4782 hCoV-19/Iceland/552/2020\|EPI_ISL_424572\|2020-03-26 | ···················· | ···················A | ···················· | ···················· | ···················· | ···················· | 1 |
| A | 4783 hCoV-19/Iceland/553/2020\|EPI_ISL_424573\|2020-03-27 | ···················· | ···················· | ···················· | ···················· | ···················· | ···················· | |
| A | 4784 hCoV-19/Iceland/554/2020\|EPI_ISL_424574\|2020-03-27 | ···················· | ···················· | ···················· | ···················· | ···················· | ···················· | |
| A | 4785 hCoV-19/Iceland/555/2020\|EPI_ISL_424575\|2020-03-28 | ···················· | ···················· | ···················· | ···················· | ···················· | ···················· | |
| A | 4786 hCoV-19/Iceland/556/2020\|EPI_ISL_424576\|2020-03-27 | ···················· | ···················· | ···················· | ···················· | ···················· | ···················· | |
| A | 4787 hCoV-19/Iceland/557/2020\|EPI_ISL_424577\|2020-03-27 | ···················· | ···················· | ···················· | ···················· | ···················· | ···················· | |
| A | 4788 hCoV-19/Iceland/558/2020\|EPI_ISL_424578\|2020-03-27 | ···················· | ···················· | ···················· | ···················· | ···················· | ···················· | |
| A | 4789 hCoV-19/Scotland/EDB017/2020\|EPI_ISL_425823\|2020-0 | ···················· | ···················· | ···················· | ···················· | ···················· | ···················· | |
| A | 4791 hCoV-19/Scotland/EDB019/2020\|EPI_ISL_425825\|2020-0 | ···················· | ···················· | ···················· | ···················· | ···················· | ···················· | |
| A | 4792 hCoV-19/Scotland/EDB020/2020\|EPI_ISL_425826\|2020-0 | ···················· | ···················· | ···················· | ···················· | ···················· | ···················· | |
| A | 4793 hCoV-19/Scotland/EDB021/2020\|EPI_ISL_425827\|2020-0 | ···················· | ···················· | ···················· | ···················· | ···················· | ···················· | |
| A | 4794 hCoV-19/Scotland/EDB022/2020\|EPI_ISL_425828\|2020-0 | ···················· | ···················· | ···················· | ···················· | ···················· | ···················· | |
| A | 4795 hCoV-19/Scotland/EDB023/2020\|EPI_ISL_425829\|2020-0 | ···················· | ···················· | ···················· | ···················· | ···················· | N··················· | |
| A | 4796 hCoV-19/Scotland/EDB039/2020\|EPI_ISL_425845\|2020-0 | ···················· | ···················· | ···················· | ···················· | ···················· | ···················· | |
| A | 4798 hCoV-19/Scotland/EDB042/2020\|EPI_ISL_425847\|2020-0 | ···················· | ···················· | ···················· | ···················· | ···················· | ···················· | |
| A | 4800 hCoV-19/Scotland/EDB045/2020\|EPI_ISL_425849\|2020-0 | ···················· | ···················· | ···················· | ···················· | ··············N····· | ···················· | |
| A | 4801 hCoV-19/Scotland/EDB046/2020\|EPI_ISL_425850\|2020-0 | ···················· | ···················· | ···················· | ···················· | ···················· | ···················· | |
| A | 4802 hCoV-19/Scotland/EDB047/2020\|EPI_ISL_425851\|2020-0 | ···················· | ···················· | ···················· | ···················· | ···················· | ···················· | |
| A | 4803 hCoV-19/Scotland/EDB048/2020\|EPI_ISL_425852\|2020-0 | ···················· | ···················· | ···················· | ···················· | ···················· | ···················· | |
| A | 4805 hCoV-19/Scotland/EDB052/2020\|EPI_ISL_425856\|2020-0 | ···················· | ···················· | ···················· | ···················· | ···················· | NN·····N············ | 1 |
| A | 4807 hCoV-19/Scotland/EDB057/2020\|EPI_ISL_425861\|2020-0 | ···················· | ···················· | ···················· | ···················· | ···················· | ···················· | |
| A | 4808 hCoV-19/Scotland/EDB058/2020\|EPI_ISL_425862\|2020-0 | ···················· | ···················· | ···················· | ···················· | ···················· | ···················· | |
| A | 4809 hCoV-19/Scotland/EDB063/2020\|EPI_ISL_425863\|2020-0 | ···················· | ···················· | ···················· | ···················· | ···················· | ···················· | |
| A | 4810 hCoV-19/Scotland/EDB065/2020\|EPI_ISL_425864\|2020-0 | ···················· | ···················· | ···················· | ···················· | ···················· | ···················· | |
| A | 4812 hCoV-19/Scotland/EDB067/2020\|EPI_ISL_425866\|2020-0 | ···················· | ···················· | ···················· | ···················· | ···················· | ···················· | |
| A | 4813 hCoV-19/Scotland/EDB068/2020\|EPI_ISL_425867\|2020-0 | ···················· | ···················· | ···················· | ···················· | ···················· | ···················· | |
| A | 4814 hCoV-19/Scotland/EDB070/2020\|EPI_ISL_425868\|2020-0 | ···················· | ···················· | ···················· | ···················· | ···················· | ···················· | |
| A | 4815 hCoV-19/Scotland/EDB071/2020\|EPI_ISL_425869\|2020-0 | ···················· | ···················· | ···················· | ···················· | ··············N····· | NN·················· | |
| A | 4816 hCoV-19/Scotland/EDB073/2020\|EPI_ISL_425870\|2020-0 | ···················· | ···················· | ···················· | ···················· | ···················· | ···················· | |
| A | 4818 hCoV-19/Scotland/EDB075/2020\|EPI_ISL_425872\|2020-0 | ···················· | ···················· | ···················· | ···················· | ···················· | ···················· | |
| A | 4819 hCoV-19/Scotland/EDB081/2020\|EPI_ISL_425873\|2020-0 | ···················· | ···················· | ···················· | ···················· | ···················· | ···················· | |
| A | 4821 hCoV-19/Scotland/EDB083/2020\|EPI_ISL_425875\|2020-0 | ···················· | ···················· | ···················· | ···················· | ···················· | ···················· | |
| A | 4822 hCoV-19/Scotland/EDB084/2020\|EPI_ISL_425876\|2020-0 | ···················· | ···················· | ···················· | ···················· | ···················· | ···················· | |
| A | 4823 hCoV-19/Scotland/EDB085/2020\|EPI_ISL_425877\|2020-0 | ···················· | ·A·················· | ···················· | ···················· | ···················· | ···················· | |
| A | 4824 hCoV-19/Scotland/EDB086/2020\|EPI_ISL_425878\|2020-0 | ···················· | ···················· | ···················· | ···················· | ···················· | ···················· | |
| A | 4825 hCoV-19/Scotland/EDB087/2020\|EPI_ISL_425879\|2020-0 | ···················· | ···················· | ···················· | ···················· | ···················· | ···················· | |
| A | 4826 hCoV-19/Scotland/EDB089/2020\|EPI_ISL_425880\|2020-0 | ···················· | ···················· | ···················· | ···················· | ···················· | ···················· | |
| A | 4827 hCoV-19/Scotland/EDB090/2020\|EPI_ISL_425881\|2020-0 | ···················· | ···················· | ···················· | ···················· | ···················· | ···················· | |
| A | 4828 hCoV-19/Scotland/EDB091/2020\|EPI_ISL_425882\|2020-0 | ···················· | ···················· | ···················· | ···················· | ···················· | ···················· | |
| A | 4831 hCoV-19/Scotland/EDB095/2020\|EPI_ISL_425885\|2020-0 | ···················· | ···················· | ···················· | ···················· | ···················· | ···················· | |
| A | 4833 hCoV-19/Scotland/EDB097/2020\|EPI_ISL_425887\|2020-0 | ···················· | ···················· | ···················· | ···················· | ···················· | ···················· | |
| A | 4834 hCoV-19/Scotland/EDB098/2020\|EPI_ISL_425888\|2020-0 | ···················· | ···················· | ···················· | ···················· | ···················· | ···················· | |
| A | 4835 hCoV-19/Scotland/EDB099/2020\|EPI_ISL_425889\|2020-0 | ···················· | ···················· | ···················· | ···················· | ···················· | ···················· | |
| A | 4837 hCoV-19/Scotland/EDB101/2020\|EPI_ISL_425891\|2020-0 | ···················· | ···················· | ···················· | ···················· | ···················· | ···················· | |
| A | 4838 hCoV-19/Scotland/EDB102/2020\|EPI_ISL_425892\|2020-0 | ···················· | ···················· | ···················· | ···················· | ···················· | ···················· | |
| A | 4839 hCoV-19/Scotland/EDB103/2020\|EPI_ISL_425893\|2020-0 | ···················· | ···················· | ···················· | ···················· | ···················· | ···················· | |
| A | 4842 hCoV-19/Scotland/EDB108/2020\|EPI_ISL_425896\|2020-0 | ···················· | ···················· | ···················· | ···················· | ···················· | ···················· | |
| A | 4843 hCoV-19/Scotland/EDB109/2020\|EPI_ISL_425897\|2020-0 | ···················· | ···················· | ···················· | ···················· | ···················· | ···················· | |
| A | 4845 hCoV-19/Scotland/EDB111/2020\|EPI_ISL_425899\|2020-0 | ···················· | ···················· | ···················· | ···················· | ···················· | ···················· | |
| A | 4846 hCoV-19/USA/WA-UW-2074/2020\|EPI_ISL_426080\|2020-03 | ···················· | ···················· | ···················· | ···················· | ···················· | ···················· | |
| A | 4847 hCoV-19/USA/WA-UW-2078/2020\|EPI_ISL_426081\|2020-03 | ···················· | ···················· | ···················· | ···················· | ···················· | ···················· | |
| A | 4848 hCoV-19/USA/WA-UW-2082/2020\|EPI_ISL_426082\|2020-03 | ···················· | ···················· | ···················· | ···················· | ···················· | ···················· | |
| A | 4849 hCoV-19/USA/WA-UW-2087/2020\|EPI_ISL_426083\|2020-03 | ···················· | ···················· | ···················· | ···················· | ···················· | ···················· | |
| A | 4850 hCoV-19/USA/WA-UW-2091/2020\|EPI_ISL_426084\|2020-03 | ···················· | ···················· | ···················· | ···················· | ···················· | ···················· | |
| A | 4851 hCoV-19/USA/WA-UW-2094/2020\|EPI_ISL_426085\|2020-03 | ···················· | ···················· | ···················· | ···················· | ···················· | ···················· | |
| A | 4852 hCoV-19/USA/WA-UW-2100/2020\|EPI_ISL_426086\|2020-03 | ···················· | ···················· | ···················· | ···················· | ···················· | ···················· | |
| A | 4853 hCoV-19/USA/WA-UW-2104/2020\|EPI_ISL_426087\|2020-03 | ···················· | ···················· | ···················· | ···················· | ···················· | ···················· | |
| A | 4854 hCoV-19/USA/WA-UW-2108/2020\|EPI_ISL_426089\|2020-03 | ···················· | ···················· | ···················· | ···················· | ···················· | ···················· | |
| A | 4855 hCoV-19/USA/WA-UW-2112/2020\|EPI_ISL_426090\|2020-03 | ···················· | ···················· | ···················· | ···················· | ···················· | ···················· | |
| A | 4856 hCoV-19/USA/WA-UW-2119/2020\|EPI_ISL_426091\|2020-03 | ···················· | ···················· | ···················· | ···················· | ···················· | ···················· | |
| A | 4857 hCoV-19/USA/WA-UW-2131/2020\|EPI_ISL_426092\|2020-03 | ···················· | ···················· | ···················· | ···················· | ···················· | ···················· | |
| A | 4858 hCoV-19/USA/WA-UW-2132/2020\|EPI_ISL_426093\|2020-03 | ···················· | ···················· | ···················· | ···················· | ···················· | ···················· | |
| A | 4859 hCoV-19/USA/WA-UW-2137/2020\|EPI_ISL_426094\|2020-03 | ···················· | ···················· | ···················· | ···················· | ···················· | ···················· | |

| | | | | | | | | |
|---|---|---|---|---|---|---|---|---|
| A | 4860 | hCoV-19/USA/WA-UW-2140/2020\|EPI_ISL_426095\|2020-03 | .................... | .................... | .................... | .................... | .................... | .................... |
| A | 4861 | hCoV-19/USA/WA-UW-2141/2020\|EPI_ISL_426096\|2020-03 | .................... | .................... | .................... | .................... | .................... | .................... |
| A | 4862 | hCoV-19/USA/WA-UW-2142/2020\|EPI_ISL_426097\|2020-03 | .................... | .................... | .................... | .................... | .................... | .................... |
| A | 4863 | hCoV-19/USA/WA-UW-2144/2020\|EPI_ISL_426098\|2020-03 | .................... | .................... | .................... | .................... | .................... | .................... |
| A | 4864 | hCoV-19/USA/WA-UW-2145/2020\|EPI_ISL_426099\|2020-03 | .................... | .................... | .................... | .................... | .................... | .................... |
| A | 4865 | hCoV-19/Switzerland/GE-SNRCI-29945373/2020\|EPI_ISL | .................... | .................... | .................... | .................... | .................... | .................... |
| A | 4866 | hCoV-19/Switzerland/BL-SNRCI-29950902/2020\|EPI_ISL | .................... | .................... | .................... | .................... | .................... | .................... |
| A | 4867 | hCoV-19/Switzerland/GE-SNRCI-29949586/2020\|EPI_ISL | .................... | .................... | .................... | .................... | .................... | .................... |
| A | 4868 | hCoV-19/Switzerland/VD-SNRCI-29965615/2020\|EPI_ISL | .................... | .................... | .................... | .................... | .................... | .................... |
| A | 4869 | hCoV-19/Scotland/CVR05/2020\|EPI_ISL_414027\|2020-03 | .................... | .................... | .................... | .................... | .................... | .................... |
| A | 4870 | hCoV-19/England/200641094/2020\|EPI_ISL_414040\|2020 | .................... | .................... | .................... | .................... | .................... | .................... |
| A | 4871 | hCoV-19/England/200690245/2020\|EPI_ISL_414041\|2020 | .................... | .................... | .................... | .................... | .................... | .................... |
| A | 4872 | hCoV-19/England/200690300/2020\|EPI_ISL_414042\|2020 | .................... | .................... | .................... | .................... | .................... | .................... |
| A | 4873 | hCoV-19/England/200690306/2020\|EPI_ISL_414043\|2020 | .................... | .................... | .................... | .................... | .................... | .................... |
| A | 4874 | hCoV-19/England/200690756/2020\|EPI_ISL_414044\|2020 | .................... | .................... | .................... | .................... | .................... | .................... |
| A | 4875 | hCoV-19/Brazil/RJ-314/2020\|EPI_ISL_414045\|2020-03- | .................... | .................... | .................... | .................... | .................... | .................... |
| A | 4876 | hCoV-19/Japan/DP0438/2020\|EPI_ISL_416600\|2020-02-1 | .................... | .................... | .................... | .................... | .................... | .................... |
| A | 4877 | hCoV-19/Japan/DP0457/2020\|EPI_ISL_416601\|2020-02-1 | .................... | .................... | .................... | .................... | .................... | .................... |
| A | 4878 | hCoV-19/Japan/DP0462/2020\|EPI_ISL_416602\|2020-02-1 | .................... | .................... | .................... | .................... | .................... | .................... |
| A | 4879 | hCoV-19/Japan/DP0464/2020\|EPI_ISL_416603\|2020-02-1 | .................... | .................... | .................... | .................... | .................... | .................... |
| A | 4880 | hCoV-19/Japan/DP0476/2020\|EPI_ISL_416604\|2020-02-1 | .................... | .................... | .................... | .................... | .................... | .................... |
| A | 4881 | hCoV-19/Japan/DP0481/2020\|EPI_ISL_416605\|2020-02-1 | .................... | .................... | .................... | .................... | .................... | .................... |
| A | 4882 | hCoV-19/Japan/DP0482/2020\|EPI_ISL_416606\|2020-02-1 | .................... | .................... | .................... | .................... | .................... | .................... |
| A | 4883 | hCoV-19/Japan/DP0543/2020\|EPI_ISL_416607\|2020-02-1 | .................... | .................... | .................... | .................... | .................... | .................... |
| A | 4884 | hCoV-19/Japan/DP0544/2020\|EPI_ISL_416608\|2020-02-1 | .................... | .................... | .................... | .................... | .................... | .................... |
| A | 4885 | hCoV-19/Japan/DP0568/2020\|EPI_ISL_416609\|2020-02-1 | .................... | .................... | .................... | .................... | .................... | .................... |
| A | 4886 | hCoV-19/Japan/DP0588/2020\|EPI_ISL_416610\|2020-02-1 | .................... | .................... | .................... | .................... | .................... | .................... |
| A | 4887 | hCoV-19/Japan/DP0644/2020\|EPI_ISL_416611\|2020-02-1 | .................... | .................... | .................... | .................... | .................... | .................... |
| A | 4888 | hCoV-19/Japan/DP0645/2020\|EPI_ISL_416612\|2020-02-1 | .................... | .................... | .................... | .................... | .................... | .................... |
| A | 4889 | hCoV-19/Japan/DP0654/2020\|EPI_ISL_416613\|2020-02-1 | .................... | .................... | .................... | .................... | .................... | .................... |
| A | 4890 | hCoV-19/Japan/DP0687/2020\|EPI_ISL_416614\|2020-02-1 | .................... | .................... | .................... | .................... | .................... | .................... |
| A | 4891 | hCoV-19/Japan/DP0690/2020\|EPI_ISL_416615\|2020-02-1 | .................... | .................... | .................... | .................... | .................... | .................... |
| A | 4892 | hCoV-19/Japan/DP0803/2020\|EPI_ISL_416630\|2020-02-1 | .................... | .................... | .................... | .................... | .................... | .................... |
| A | 4893 | hCoV-19/Japan/DP0804/2020\|EPI_ISL_416631\|2020-02-1 | .................... | .................... | .................... | .................... | .................... | .................... |
| A | 4894 | hCoV-19/Japan/DP0827/2020\|EPI_ISL_416632\|2020-02-1 | .................... | .................... | .................... | .................... | .................... | .................... |
| A | 4895 | hCoV-19/Japan/DP0880/2020\|EPI_ISL_416633\|2020-02-1 | .................... | .................... | .................... | .................... | .................... | .................... |
| A | 4896 | hCoV-19/USA/WA-UW98/2020\|EPI_ISL_416636\|2020-03-12 | .................... | .................... | .................... | .................... | .................... | .................... |
| A | 4897 | hCoV-19/USA/WA-UW99/2020\|EPI_ISL_416637\|2020-03-12 | .................... | .................... | .................... | .................... | .................... | .................... |
| A | 4898 | hCoV-19/USA/WA-UW142/2020\|EPI_ISL_416680\|2020-03-1 | .................... | .................... | .................... | .................... | .................... | .................... |
| A | 4899 | hCoV-19/USA/WA-UW143/2020\|EPI_ISL_416681\|2020-03-1 | .................... | .................... | .................... | .................... | .................... | .................... |
| A | 4900 | hCoV-19/USA/WA-UW144/2020\|EPI_ISL_416682\|2020-03-0 | .................... | .................... | .................... | .................... | .................... | .................... |
| A | 4901 | hCoV-19/USA/WA-UW145/2020\|EPI_ISL_416683\|2020-03-1 | .................... | .................... | .................... | .................... | .................... | .................... |
| A | 4902 | hCoV-19/USA/WA-UW146/2020\|EPI_ISL_416684\|2020-03-1 | .................... | .................... | .................... | .................... | .................... | .................... |
| A | 4903 | hCoV-19/USA/WA-UW147/2020\|EPI_ISL_416685\|2020-03-1 | .................... | .................... | .................... | .................... | .................... | .................... |
| A | 4904 | hCoV-19/USA/WA-UW148/2020\|EPI_ISL_416686\|2020-03-1 | .................... | .................... | .................... | .................... | .................... | .................... |
| A | 4905 | hCoV-19/USA/WA-UW149/2020\|EPI_ISL_416687\|2020-03-1 | .................... | .................... | .................... | .................... | .................... | .................... |
| A | 4906 | hCoV-19/USA/WA-UW150/2020\|EPI_ISL_416688\|2020-03-1 | .................... | .................... | .................... | .................... | .................... | .................... |
| A | 4907 | hCoV-19/USA/WA-UW151/2020\|EPI_ISL_416689\|2020-03-1 | .................... | .................... | .................... | .................... | .................... | .................... |
| A | 4908 | hCoV-19/USA/WA-UW152/2020\|EPI_ISL_416690\|2020-03-1 | .................... | .................... | .................... | .................... | .................... | .................... |
| A | 4909 | hCoV-19/USA/WA-UW153/2020\|EPI_ISL_416691\|2020-03-1 | .................... | .................... | .................... | .................... | .................... | .................... |
| A | 4910 | hCoV-19/USA/WA-UW154/2020\|EPI_ISL_416692\|2020-03-1 | .................... | .................... | .................... | .................... | .................... | .................... |
| A | 4911 | hCoV-19/USA/WA-UW155/2020\|EPI_ISL_416693\|2020-03-1 | .................... | .................... | .................... | .................... | .................... | .................... |
| A | 4912 | hCoV-19/USA/WA-UW156/2020\|EPI_ISL_416694\|2020-03-1 | .................... | .................... | .................... | .................... | .................... | .................... |
| A | 4913 | hCoV-19/USA/CT-UW158/2020\|EPI_ISL_416696\|2020-03-1 | .................... | .................... | .................... | .................... | .................... | .................... |
| A | 4914 | hCoV-19/USA/CT-UW159/2020\|EPI_ISL_416697\|2020-03-1 | .................... | .................... | .................... | .................... | .................... | .................... |
| A | 4915 | hCoV-19/USA/WA-UW160/2020\|EPI_ISL_416698\|2020-03-1 | .................... | .................... | .................... | .................... | .................... | .................... |
| A | 4916 | hCoV-19/USA/WA-UW161/2020\|EPI_ISL_416699\|2020-03-1 | .................... | .................... | .................... | .................... | .................... | .................... |
| A | 4917 | hCoV-19/USA/CA-CZB05/2020\|EPI_ISL_417931\|2020-03-1 | .................... | .................... | .................... | .................... | .................... | .................... |
| A | 4918 | hCoV-19/USA/CA-CZB06/2020\|EPI_ISL_417932\|2020-03-1 | .................... | .................... | .................... | .................... | .................... | .................... |
| A | 4919 | hCoV-19/USA/CA-CZB07/2020\|EPI_ISL_417933\|2020-03-1 | .................... | .................... | .................... | .................... | .................... | .................... |
| A | 4920 | hCoV-19/USA/CA-CZB011a/2020\|EPI_ISL_417935\|2020-03 | .................... | .................... | .................... | .................... | .................... | .................... |
| A | 4921 | hCoV-19/USA/CA-CZB013/2020\|EPI_ISL_417937\|2020-03- | .................... | .................... | .................... | .................... | .................... | .................... |
| A | 4922 | hCoV-19/USA/CA-CZB014/2020\|EPI_ISL_417938\|2020-03- | .................... | .................... | .................... | .................... | .................... | .................... |
| A | 4923 | hCoV-19/USA/CA-CZB015a/2020\|EPI_ISL_417939\|2020-03 | .................... | .................... | .................... | .................... | .................... | .................... |
| A | 4924 | hCoV-19/DRC/73/2020\|EPI_ISL_417941\|2020-03-18 | .................... | .................... | .................... | .................... | .................... | .................... |
| A | 4925 | hCoV-19/DRC/80/2020\|EPI_ISL_417942\|2020-03-18 | .................... | .................... | .................... | .................... | .................... | .................... |
| A | 4926 | hCoV-19/DRC/81/2020\|EPI_ISL_417944\|2020-03-18 | .................... | .................... | .................... | .................... | .................... | .................... |
| A | 4927 | hCoV-19/DRC/82/2020\|EPI_ISL_417946\|2020-03-18 | .................... | .................... | .................... | .................... | .................... | .................... |
| A | 4928 | hCoV-19/USA/UT-012/2020\|EPI_ISL_417960\|2020-03-12 | .................... | .................... | .................... | .................... | .................... | .................... |
| A | 4929 | hCoV-19/Spain/MD-HLP-H9-38/2020\|EPI_ISL_417961\|202 | .................... | .................... | .................... | .................... | .................... | .................... |
| A | 4930 | hCoV-19/Spain/MD-HLP-H10-39/2020\|EPI_ISL_417963\|20 | .................... | .................... | .................... | .................... | .................... | .................... |
| A | 4931 | hCoV-19/USA/UT-014/2020\|EPI_ISL_417964\|2020-03-13 | .................... | .................... | .................... | .................... | .................... | .................... |
| A | 4932 | hCoV-19/Spain/MD-HLP-H11-40/2020\|EPI_ISL_417967\|20 | .................... | .................... | .................... | .................... | .................... | .................... |
| A | 4933 | hCoV-19/Belgium/ULG-10012/2020\|EPI_ISL_421190\|2020 | .................... | .................... | .................... | .................... | .................... | .................... |
| A | 4934 | hCoV-19/Belgium/ULG-10013/2020\|EPI_ISL_421191\|2020 | .................... | .................... | .................... | .................... | .................... | .................... |
| A | 4935 | hCoV-19/Belgium/ULG-10014/2020\|EPI_ISL_421192\|2020 | .................... | .................... | .................... | .................... | .................... | .................... |
| A | 4936 | hCoV-19/Belgium/ULG-10015/2020\|EPI_ISL_421193\|2020 | .................... | .................... | .................... | .................... | .................... | .................... |
| A | 4937 | hCoV-19/Belgium/ULG-10016/2020\|EPI_ISL_421194\|2020 | .................... | .................... | .................... | .................... | .................... | .................... |

| | | | | | | | | |
|---|---|---|---|---|---|---|---|---|
| A | 4938 | hCoV-19/Belgium/ULG-10017/2020\|EPI_ISL_421195\|2020 | ···················· | ···················· | ····················· | ···················· | ···················· | ···················· |
| A | 4939 | hCoV-19/Belgium/ULG-10018/2020\|EPI_ISL_421196\|2020 | ···················· | ···················· | ····················· | ···················· | ···················· | ···················· |
| A | 4940 | hCoV-19/Belgium/ULG-10024/2020\|EPI_ISL_421198\|2020 | ···················· | ···················· | ····················· | ···················· | ···················· | ···················· |
| A | 4941 | hCoV-19/Belgium/ULG-10029/2020\|EPI_ISL_421203\|2020 | ···················· | ···················· | ····················· | ···················· | ···················· | ···················· |
| A | 4942 | hCoV-19/Belgium/ULG-10030/2020\|EPI_ISL_421204\|2020 | ···················· | ···················· | ····················· | ···················· | ···················· | ···················· |
| A | 4943 | hCoV-19/Belgium/ULG-10031/2020\|EPI_ISL_421205\|2020 | ···················· | ···················· | ····················· | ···················· | ···················· | ···················· |
| A | 4944 | hCoV-19/Belgium/ULG-10032/2020\|EPI_ISL_421206\|2020 | ···················· | ···················· | ····················· | ···················· | ···················· | ···················· |
| A | 4945 | hCoV-19/Belgium/ULG-10033/2020\|EPI_ISL_421207\|2020 | ···················· | ···················· | ····················· | ···················· | ···················· | ···················· |
| A | 4946 | hCoV-19/Belgium/ULG-10043/2020\|EPI_ISL_421210\|2020 | ···················· | ···················· | ····················· | ···················· | ···················· | ···················· |
| A | 4947 | hCoV-19/Belgium/ULG-10045/2020\|EPI_ISL_421212\|2020 | ···················· | ···················· | ····················· | ···················· | ···················· | ···················· |
| A | 4948 | hCoV-19/Belgium/ULG-10046/2020\|EPI_ISL_421213\|2020 | ···················· | ···················· | ····················· | ···················· | ···················· | ···················· |
| A | 4949 | hCoV-19/Belgium/ULG-10047/2020\|EPI_ISL_421214\|2020 | ···················· | ···················· | ····················· | ···················· | ···················· | ···················· |
| A | 4950 | hCoV-19/Hangzhou/HZCDC6706/2020\|EPI_ISL_421222\|202 | ···················· | ···················· | ····················· | ···················· | ···················· | ···················· |
| A | 4951 | hCoV-19/USA/NY-PV09172/2020\|EPI_ISL_422534\|2020-03 | ···················· | ···················· | ····················· | ···················· | ···················· | ···················· |
| A | 4952 | hCoV-19/USA/NY-PV09176/2020\|EPI_ISL_422535\|2020-03 | ···················· | ···················· | ····················· | ···················· | ···················· | ···················· |
| A | 4953 | hCoV-19/USA/NY-PV09177/2020\|EPI_ISL_422536\|2020-03 | ···················· | ···················· | ····················· | ···················· | ···················· | ···················· |
| A | 4954 | hCoV-19/USA/NY-PV09178/2020\|EPI_ISL_422537\|2020-03 | ···················· | ···················· | ····················· | ···················· | ···················· | ···················· |
| A | 4955 | hCoV-19/USA/NY-PV09180/2020\|EPI_ISL_422539\|2020-03 | ···················· | ···················· | ····················· | ···················· | ···················· | ···················· |
| A | 4956 | hCoV-19/USA/NY-PV09181/2020\|EPI_ISL_422540\|2020-03 | ···················· | ···················· | ····················· | ···················· | ···················· | ···················· |
| A | 4957 | hCoV-19/USA/NY-PV09182/2020\|EPI_ISL_422541\|2020-03 | ···················· | ···················· | ····················· | ···················· | ···················· | ···················· |
| A | 4958 | hCoV-19/USA/NY-PV09183/2020\|EPI_ISL_422542\|2020-03 | ···················· | ···················· | ····················· | ···················· | ···················· | ···················· |
| A | 4959 | hCoV-19/USA/NY-PV09186/2020\|EPI_ISL_422543\|2020-03 | ···················· | ···················· | ····················· | ···················· | ···················· | ···················· |
| A | 4960 | hCoV-19/USA/NY-PV09187/2020\|EPI_ISL_422544\|2020-03 | ···················· | ···················· | ····················· | ···················· | ···················· | ···················· |
| A | 4961 | hCoV-19/USA/NY-PV09189/2020\|EPI_ISL_422546\|2020-03 | ···················· | ·······C············ | ····················· | ···················· | ···················· | ···················· |
| A | 4962 | hCoV-19/USA/NY-PV09190/2020\|EPI_ISL_422547\|2020-03 | ···················· | ···················· | ····················· | ···················· | ···················· | ···················· |
| A | 4963 | hCoV-19/USA/NY-PV09192/2020\|EPI_ISL_422548\|2020-03 | ···················· | ···················· | ····················· | ···················· | ···················· | ···················· |
| A | 4964 | hCoV-19/USA/NY-PV09193/2020\|EPI_ISL_422549\|2020-03 | ···················· | ···················· | ····················· | ···················· | ···················· | ···················· |
| A | 4965 | hCoV-19/USA/NY-PV09194/2020\|EPI_ISL_422550\|2020-03 | ···················· | ···················· | ····················· | ···················· | ···················· | ···················· |
| A | 4966 | hCoV-19/USA/NY-PV09195/2020\|EPI_ISL_422551\|2020-03 | ···················· | ···················· | ····················· | ···················· | ···················· | ···················· |
| A | 4967 | hCoV-19/England/20126001604/2020\|EPI_ISL_423865\|20 | ···················· | ·A·················· | ····················· | ···················· | ···················· | ···················· |
| A | 4969 | hCoV-19/England/20126001904/2020\|EPI_ISL_423867\|20 | ···················· | ···················· | ····················· | ···················· | ···················· | ···················· |
| A | 4970 | hCoV-19/England/20126003406/2020\|EPI_ISL_423877\|20 | ···················· | ···················· | ····················· | ···················· | ···················· | ···················· |
| A | 4971 | hCoV-19/Scotland/EDB407/2020\|EPI_ISL_426000\|2020-0 | ···················· | ···················· | ····················· | ···················· | ···················· | ···················· |
| A | 4972 | hCoV-19/Scotland/EDB409/2020\|EPI_ISL_426001\|2020-0 | ···················· | ···················· | ····················· | ···················· | ···················· | ···················· |
| A | 4973 | hCoV-19/Scotland/EDB410/2020\|EPI_ISL_426002\|2020-0 | ···················· | ···················· | ····················· | ···················· | ···················· | ···················· |
| A | 4974 | hCoV-19/Scotland/EDB412/2020\|EPI_ISL_426003\|2020-0 | ···················· | ···················· | ····················· | ···················· | ···················· | ···················· |
| A | 4975 | hCoV-19/Scotland/EDB413/2020\|EPI_ISL_426004\|2020-0 | ···················· | ···················· | ····················· | ···················· | ···················· | ···················· |
| A | 4976 | hCoV-19/Scotland/EDB420/2020\|EPI_ISL_426005\|2020-0 | ···················· | ···················· | ····················· | ···················· | ···················· | ···················· |
| A | 4977 | hCoV-19/Scotland/EDB421/2020\|EPI_ISL_426006\|2020-0 | ···················· | ···················· | ····················· | ···················· | ···················· | ···················· |
| A | 4978 | hCoV-19/Scotland/EDB442/2020\|EPI_ISL_426007\|2020-0 | ···················· | ···················· | ····················· | ···················· | ···················· | ···················· |
| A | 4979 | hCoV-19/Scotland/EDB501/2020\|EPI_ISL_426008\|2020-0 | ···················· | ···················· | ····················· | ···················· | ···················· | ···················· |
| A | 4980 | hCoV-19/Scotland/EDB502/2020\|EPI_ISL_426009\|2020-0 | ···················· | ···················· | ····················· | ···················· | ···················· | ···················· |
| A | 4981 | hCoV-19/Scotland/EDB508/2020\|EPI_ISL_426010\|2020-0 | ···················· | ···················· | ····················· | ···················· | ···················· | ···················· |
| A | 4982 | hCoV-19/Scotland/EDB513/2020\|EPI_ISL_426011\|2020-0 | ···················· | ···················· | ····················· | ···················· | ···················· | ···················· |
| A | 4983 | hCoV-19/Scotland/EDB517/2020\|EPI_ISL_426012\|2020-0 | ···················· | ···················· | ····················· | ···················· | ···················· | ···················· |
| A | 4984 | hCoV-19/Scotland/EDB530/2020\|EPI_ISL_426013\|2020-0 | ···················· | ···················· | ····················· | ···················· | ···················· | ···················· |
| A | 4985 | hCoV-19/Scotland/EDB533/2020\|EPI_ISL_426014\|2020-0 | ···················· | ···················· | ····················· | ···················· | ···················· | ···················· |
| A | 4986 | hCoV-19/Scotland/EDB551/2020\|EPI_ISL_426015\|2020-0 | ···················· | ···················· | ····················· | ···················· | ···················· | ···················· |
| A | 4987 | hCoV-19/Scotland/EDB555/2020\|EPI_ISL_426016\|2020-0 | ···················· | ···················· | ····················· | ···················· | ···················· | ···················· |
| A | 4988 | hCoV-19/Scotland/EDB557/2020\|EPI_ISL_426017\|2020-0 | ···················· | ···················· | ····················· | ···················· | ···················· | ···················· |
| A | 4989 | hCoV-19/Scotland/EDB560/2020\|EPI_ISL_426019\|2020-0 | ···················· | ···················· | ····················· | ···················· | ···················· | ···················· |
| A | 4990 | hCoV-19/Scotland/EDB561/2020\|EPI_ISL_426020\|2020-0 | ···················· | ···················· | ····················· | ···················· | ···················· | ···················· |
| A | 4991 | hCoV-19/Scotland/EDB562/2020\|EPI_ISL_426021\|2020-0 | ···················· | ·A·················· | ····················· | ···················· | ···················· | ···················· |
| A | 4992 | hCoV-19/Scotland/EDB563/2020\|EPI_ISL_426022\|2020-0 | ···················· | ···················· | ····················· | ···················· | ···················· | ···················· |
| A | 4993 | hCoV-19/Scotland/EDB565/2020\|EPI_ISL_426023\|2020-0 | ·T·················· | ···················· | ····················· | ···················· | ···················· | ···················· |
| A | 4994 | hCoV-19/Scotland/EDB566/2020\|EPI_ISL_426024\|2020-0 | ···················· | ···················· | ····················· | ···················· | ···················· | ···················· |
| A | 4995 | hCoV-19/USA/NY-Wadsworth-10496-01/2020\|EPI_ISL_426 | ···················· | ···················· | ····················· | ···················· | ···················· | ···················· |
| A | 4996 | hCoV-19/USA/NY-Wadsworth-10528-01/2020\|EPI_ISL_426 | ···················· | ···················· | ····················· | ···················· | ···················· | ···················· |
| A | 4997 | hCoV-19/USA/NY-Wadsworth-10683-01/2020\|EPI_ISL_426 | ···················· | ···················· | ····················· | ···················· | ···················· | ···················· |
| A | 4998 | hCoV-19/USA/NY-Wadsworth-10690-01/2020\|EPI_ISL_426 | ···················· | ···················· | ····················· | ···················· | ···················· | ···················· |
| A | 4999 | hCoV-19/USA/NY-Wadsworth-10695-01/2020\|EPI_ISL_426 | ···················· | ···················· | ····················· | ···················· | ···················· | ···················· |
| A | 5000 | hCoV-19/USA/NY-Wadsworth-10703-01/2020\|EPI_ISL_426 | ···················· | ···················· | ····················· | ···················· | ···················· | ···················· |
| A | 5001 | hCoV-19/USA/NY-Wadsworth-10704-01/2020\|EPI_ISL_426 | ···················· | ···················· | ····················· | ···················· | ···················· | ···················· |
| A | 5002 | hCoV-19/USA/NY-Wadsworth-10707-01/2020\|EPI_ISL_426 | ···················· | ···················· | ····················· | ···················· | ···················· | ···················· |
| A | 5003 | hCoV-19/USA/NY-Wadsworth-10935-01/2020\|EPI_ISL_426 | ···················· | ···················· | ····················· | ···················· | ···················· | ···················· |
| A | 5004 | hCoV-19/USA/NY-Wadsworth-10955-01/2020\|EPI_ISL_426 | ···················· | ···················· | ····················· | ···················· | ···················· | ···················· |
| A | 5005 | hCoV-19/USA/NY-Wadsworth-10957-02/2020\|EPI_ISL_426 | ···················· | ···················· | ····················· | ···················· | ···················· | ···················· |
| A | 5006 | hCoV-19/USA/NY-Wadsworth-10958-01/2020\|EPI_ISL_426 | ···················· | ···················· | ····················· | ···················· | ···················· | ···················· |
| A | 5007 | hCoV-19/USA/NY-Wadsworth-10999-01/2020\|EPI_ISL_426 | ···················· | ···················· | ····················· | ···················· | ···················· | ···················· |
| A | 5008 | hCoV-19/USA/NY-Wadsworth-11000-01/2020\|EPI_ISL_426 | ···················· | ···················· | ····················· | ···················· | ···················· | ···················· |
| A | 5009 | hCoV-19/USA/NY-Wadsworth-11003-01/2020\|EPI_ISL_426 | ···················· | ···················· | ····················· | ···················· | ···················· | ···················· |
| A | 5010 | hCoV-19/USA/NY-Wadsworth-11017-01/2020\|EPI_ISL_426 | ···················· | ···················· | ····················· | ···················· | ···················· | ···················· |
| A | 5011 | hCoV-19/USA/NY-Wadsworth-11180-01/2020\|EPI_ISL_426 | ···················· | ···················· | ····················· | ···················· | ···················· | ···················· |
| A | 5012 | hCoV-19/USA/NY-Wadsworth-11202-01/2020\|EPI_ISL_426 | ···················· | ···················· | ····················· | ···················· | ···················· | ···················· |
| A | 5013 | hCoV-19/USA/NY-Wadsworth-11284-01/2020\|EPI_ISL_426 | ···················· | ···················· | ····················· | ···················· | ···················· | ···················· |
| A | 5014 | hCoV-19/USA/NY-Wadsworth-11291-01/2020\|EPI_ISL_426 | ···················· | ···················· | ····················· | ···················· | ···················· | ···················· |
| A | 5015 | hCoV-19/USA/NY-Wadsworth-11344-01/2020\|EPI_ISL_426 | ···················· | ···················· | ····················· | ···················· | ···················· | ···················· |
| A | 5016 | hCoV-19/USA/NY-Wadsworth-11353-01/2020\|EPI_ISL_426 | ···················· | ···················· | ····················· | ···················· | ···················· | ···················· |

| | | | | | | | | |
|---|---|---|---|---|---|---|---|---|
| A | 5017 | hCoV-19/USA/NY-Wadsworth-11354-01/2020\|EPI_ISL_426 | .................... | .................... | .................... | .................... | .................... | .................... |
| A | 5018 | hCoV-19/USA/NY-Wadsworth-11379-01/2020\|EPI_ISL_426 | .................... | .................... | .................... | .................... | .................... | .................... |
| A | 5019 | hCoV-19/USA/NY-Wadsworth-11380-01/2020\|EPI_ISL_426 | .................... | .................... | .................... | .................... | .................... | .................... |
| A | 5020 | hCoV-19/USA/NY-Wadsworth-11666-01/2020\|EPI_ISL_426 | .................... | .................... | .................... | .................... | .................... | .................... |
| A | 5021 | hCoV-19/Czech Republic/2282/2020\|EPI_ISL_426051\|20 | .................... | .................... | .................... | .................... | .................... | .................... |
| A | 5022 | hCoV-19/USA/CT-UW-4344/2020\|EPI_ISL_426052\|2020-03 | .................... | .................... | .................... | .................... | .................... | .................... |
| A | 5023 | hCoV-19/USA/CT-UW-4346/2020\|EPI_ISL_426053\|2020-03 | .................... | .................... | .................... | .................... | .................... | .................... |
| A | 5024 | hCoV-19/USA/CT-UW-4347/2020\|EPI_ISL_426054\|2020-03 | .................... | .................... | .................... | .................... | .................... | .................... |
| A | 5025 | hCoV-19/USA/CT-UW-4366/2020\|EPI_ISL_426055\|2020-03 | .................... | .................... | .................... | .................... | .................... | .................... |
| A | 5026 | hCoV-19/USA/CT-UW-4372/2020\|EPI_ISL_426056\|2020-03 | .................... | .................... | .................... | .................... | .................... | .................... |
| A | 5027 | hCoV-19/USA/ID-UW-2127/2020\|EPI_ISL_426057\|2020-03 | .................... | .................... | .................... | .................... | .................... | .................... |
| A | 5028 | hCoV-19/USA/ID-UW-2134/2020\|EPI_ISL_426058\|2020-03 | .................... | .................... | .................... | .................... | .................... | .................... |
| A | 5029 | hCoV-19/USA/ID-UW-2159/2020\|EPI_ISL_426059\|2020-03 | .................... | .................... | .................... | .................... | .................... | .................... |
| A | 5030 | hCoV-19/USA/ID-UW-2160/2020\|EPI_ISL_426060\|2020-03 | .................... | .................... | .................... | .................... | .................... | .................... |
| A | 5031 | hCoV-19/USA/ID-UW-2174/2020\|EPI_ISL_426061\|2020-03 | .................... | .................... | .................... | .................... | .................... | .................... |
| A | 5032 | hCoV-19/USA/ID-UW-2198/2020\|EPI_ISL_426062\|2020-03 | .................... | .................... | .................... | .................... | .................... | .................... |
| A | 5033 | hCoV-19/USA/ID-UW-2255/2020\|EPI_ISL_426063\|2020-03 | .................... | .................... | .................... | .................... | .................... | .................... |
| A | 5034 | hCoV-19/USA/ID-UW-4355/2020\|EPI_ISL_426064\|2020-03 | .................... | .................... | .................... | .................... | .................... | .................... |
| A | 5035 | hCoV-19/USA/ID-UW-4357/2020\|EPI_ISL_426065\|2020-03 | .................... | .................... | .................... | .................... | .................... | .................... |
| A | 5036 | hCoV-19/USA/ID-UW-4375/2020\|EPI_ISL_426066\|2020-03 | .................... | .................... | .................... | .................... | .................... | .................... |
| A | 5037 | hCoV-19/USA/ID-UW-4378/2020\|EPI_ISL_426067\|2020-03 | .................... | .................... | .................... | .................... | .................... | .................... |
| A | 5038 | hCoV-19/USA/ID-UW-4380/2020\|EPI_ISL_426068\|2020-03 | .................... | .................... | .................... | .................... | .................... | .................... |
| A | 5039 | hCoV-19/USA/ID-UW-4396/2020\|EPI_ISL_426069\|2020-03 | .................... | .................... | .................... | .................... | .................... | .................... |
| A | 5040 | hCoV-19/USA/ID-UW-4400/2020\|EPI_ISL_426070\|2020-03 | .................... | .................... | .................... | .................... | .................... | .................... |
| A | 5041 | hCoV-19/USA/ID-UW-4427/2020\|EPI_ISL_426072\|2020-03 | .................... | .................... | .................... | .................... | .................... | .................... |
| A | 5042 | hCoV-19/USA/ID-UW-4433/2020\|EPI_ISL_426073\|2020-03 | .................... | .................... | .................... | .................... | .................... | .................... |
| A | 5043 | hCoV-19/USA/ID-UW-4448/2020\|EPI_ISL_426074\|2020-03 | .................... | .................... | .................... | .................... | .................... | .................... |
| A | 5044 | hCoV-19/USA/ID-UW-4461/2020\|EPI_ISL_426075\|2020-03 | .................... | .................... | .................... | .................... | .................... | .................... |
| A | 5045 | hCoV-19/USA/OR-UW-4388/2020\|EPI_ISL_426076\|2020-03 | .................... | .................... | .................... | .................... | .................... | .................... |
| A | 5046 | hCoV-19/USA/UN-UW-2069/2020\|EPI_ISL_426077\|2020-03 | .................... | .................... | .................... | .................... | .................... | .................... |
| A | 5047 | hCoV-19/USA/UN-UW-2139/2020\|EPI_ISL_426078\|2020-03 | .................... | .................... | .................... | .................... | .................... | .................... |
| A | 5048 | hCoV-19/USA/WA-UW-2072/2020\|EPI_ISL_426079\|2020-03 | .................... | .................... | .................... | .................... | .................... | .................... |
| A | 5049 | hCoV-19/Russia/StPetersburg-RII4711S/2020\|EPI_ISL_ | .................... | .................... | .................... | .................... | .................... | .................... |
| A | 5050 | hCoV-19/Russia/StPetersburg-RII4713S/2020\|EPI_ISL_ | .................... | .................... | .................... | .................... | .................... | .................... |
| A | 5051 | hCoV-19/Russia/StPetersburg-RII4723S/2020\|EPI_ISL_ | .................... | .................... | .................... | .................... | .................... | .................... |
| A | 5052 | hCoV-19/Russia/StPetersburg-RII4724S/2020\|EPI_ISL_ | .................... | .................... | .................... | .................... | .................... | .................... |
| A | 5053 | hCoV-19/Russia/StPetersburg-RII4726S/2020\|EPI_ISL_ | .................... | .................... | .................... | .................... | .................... | .................... |
| A | 5054 | hCoV-19/Russia/StPetersburg-RII4728S/2020\|EPI_ISL_ | .................... | .................... | .................... | .................... | .................... | .................... |
| A | 5055 | hCoV-19/Russia/StPetersburg-RII4781S/2020\|EPI_ISL_ | .................... | .................... | .................... | .................... | .................... | .................... |
| A | 5056 | hCoV-19/Russia/StPetersburg-RII4326S/2020\|EPI_ISL_ | .................... | .................... | .................... | .................... | .................... | .................... |
| A | 5057 | hCoV-19/Russia/StPetersburg-RII4328S/2020\|EPI_ISL_ | .................... | .................... | .................... | .................... | .................... | .................... |
| A | 5058 | hCoV-19/Russia/StPetersburg-RII4332S/2020\|EPI_ISL_ | .................... | .................... | .................... | .................... | .................... | .................... |
| A | 5059 | hCoV-19/Belgium/ULG-10098/2020\|EPI_ISL_427349\|2020 | .................... | .................... | .................... | .................... | .................... | .................... |
| A | 5060 | hCoV-19/Belgium/ULG-10099/2020\|EPI_ISL_427350\|2020 | .................... | .................... | .................... | .................... | .................... | .................... |
| A | 5061 | hCoV-19/Belgium/ULG-10100/2020\|EPI_ISL_427351\|2020 | .................... | .................... | .................... | .................... | .................... | .................... |
| A | 5062 | hCoV-19/Belgium/ULG-10101/2020\|EPI_ISL_427352\|2020 | .................... | .................... | .................... | .................... | .................... | .................... |
| A | 5063 | hCoV-19/Belgium/ULG-10103/2020\|EPI_ISL_427354\|2020 | .................... | .................... | .................... | .................... | .................... | .................... |
| A | 5064 | hCoV-19/Belgium/ULG-10106/2020\|EPI_ISL_427357\|2020 | .................... | .................... | .................... | .................... | .................... | .................... |
| A | 5065 | hCoV-19/Belgium/ULG-10107/2020\|EPI_ISL_427358\|2020 | .................... | .................... | .................... | .................... | .................... | .................... |
| A | 5066 | hCoV-19/Belgium/ULG-10108/2020\|EPI_ISL_427359\|2020 | .................... | .................... | .................... | .................... | .................... | .................... |
| A | 5067 | hCoV-19/Belgium/ULG-10115/2020\|EPI_ISL_427364\|2020 | .................... | .................... | .................... | .................... | .................... | .................... |
| A | 5068 | hCoV-19/Belgium/ULG-10118/2020\|EPI_ISL_427366\|2020 | .................... | .................... | .................... | .................... | .................... | .................... |
| A | 5069 | hCoV-19/Belgium/ULG-10122/2020\|EPI_ISL_427370\|2020 | .................... | .................... | .................... | .................... | .................... | .................... |
| A | 5070 | hCoV-19/Belgium/ULG-10124/2020\|EPI_ISL_427371\|2020 | .................... | .................... | .................... | .................... | .................... | .................... |
| A | 5071 | hCoV-19/Belgium/ULG-10126/2020\|EPI_ISL_427372\|2020 | .................... | .................... | .................... | .................... | .................... | .................... |
| A | 5072 | hCoV-19/Belgium/ULG-10134/2020\|EPI_ISL_427379\|2020 | .................... | .................... | .................... | .................... | .................... | .................... |
| A | 5073 | hCoV-19/Belgium/ULG-10142/2020\|EPI_ISL_427385\|2020 | .................... | .................... | .................... | .................... | .................... | .................... |
| A | 5074 | hCoV-19/Belgium/ULG-10144/2020\|EPI_ISL_427388\|2020 | .................... | .................... | .................... | .................... | .................... | .................... |
| A | 5075 | hCoV-19/Belgium/ULG-10125/2020\|EPI_ISL_427390\|2020 | .................... | .................... | .................... | .................... | .................... | .................... |
| A | 5076 | hCoV-19/Turkey/IST-GLAB-008/2020\|EPI_ISL_427391\|20 | .................... | .................... | .................... | .................... | .................... | .................... |
| A | 5077 | hCoV-19/Taiwan/TSGH-05/2020\|EPI_ISL_427392\|2020-03 | .................... | .................... | .................... | .................... | .................... | .................... |
| A | 5078 | hCoV-19/Taiwan/TSGH-06/2020\|EPI_ISL_427393\|2020-03 | .................... | .................... | .................... | .................... | .................... | .................... |
| A | 5079 | hCoV-19/Taiwan/TSGH-07/2020\|EPI_ISL_427394\|2020-03 | .................... | .................... | .................... | .................... | .................... | .................... |
| A | 5080 | hCoV-19/Taiwan/TSGH-08/2020\|EPI_ISL_427395\|2020-03 | .................... | .................... | .................... | .................... | .................... | .................... |
| A | 5081 | hCoV-19/Taiwan/TSGH-11/2020\|EPI_ISL_427398\|2020-03 | .................... | .................... | .................... | .................... | .................... | .................... |
| A | 5082 | hCoV-19/Sri Lanka/COV38/2020\|EPI_ISL_428670\|2020-0 | .................... | .................... | .................... | .................... | .................... | .................... |
| A | 5083 | hCoV-19/Sri Lanka/COV53/2020\|EPI_ISL_428671\|2020-0 | .................... | .................... | .................... | .................... | .................... | .................... |
| A | 5084 | hCoV-19/Sri Lanka/COV91/2020\|EPI_ISL_428672\|2020-0 | .................... | .................... | .................... | .................... | .................... | .................... |
| A | 5085 | hCoV-19/Sri Lanka/COV486/2020\|EPI_ISL_428673\|2020- | .................... | .................... | .................... | .................... | .................... | .................... |
| A | 5086 | hCoV-19/Spain/MD-H12-LP11-2271/2020\|EPI_ISL_428674 | .................... | .................... | .................... | .................... | .................... | .................... |
| A | 5087 | hCoV-19/Spain/MD-H12-LP16-6193/2020\|EPI_ISL_428675 | .................... | .................... | .................... | .................... | .................... | .................... |
| A | 5088 | hCoV-19/Spain/MD-H12-LP19-4952/2020\|EPI_ISL_428676 | .................... | .................... | .................... | .................... | .................... | .................... |
| A | 5089 | hCoV-19/Spain/MD-H12-LP20-2327/2020\|EPI_ISL_428677 | .................... | .................... | .................... | .................... | .................... | .................... |
| A | 5090 | hCoV-19/Spain/MD-H12-LP22-5885/2020\|EPI_ISL_428678 | .................... | .................... | .................... | .................... | .................... | .................... |
| A | 5091 | hCoV-19/Spain/MD-H12-LP23-5852/2020\|EPI_ISL_428679 | .................... | .................... | .................... | .................... | .................... | .................... |
| A | 5092 | hCoV-19/Spain/MD-H12-LP24-5999/2020\|EPI_ISL_428680 | .................... | .................... | .................... | .................... | .................... | .................... |
| A | 5093 | hCoV-19/Spain/MD-H12-LP27-2548/2020\|EPI_ISL_428681 | .................... | .................... | .................... | .................... | .................... | .................... |
| A | 5094 | hCoV-19/Spain/MD-H12-LP30-2226/2020\|EPI_ISL_428682 | .................... | .................... | .................... | .................... | .................... | .................... |

| | | | |
|---|---|---|---|
| A | 5095 | hCoV-19/Spain/MD-H12-13/2020\|EPI_ISL_428683\|2020-0 | ···················· ···················· ····················· ···················· ···················· ···················· |
| A | 5096 | hCoV-19/Spain/MD-H12-20/2020\|EPI_ISL_428684\|2020-0 | ···················· ···················· ····················· ···················· ···················· ···················· |
| A | 5097 | hCoV-19/Spain/MD-H12-23/2020\|EPI_ISL_428686\|2020-0 | ···················· ···················· ····················· ···················· ···················· ···················· |
| A | 5098 | hCoV-19/Spain/MD-H12-24/2020\|EPI_ISL_428687\|2020-0 | ···················· ···················· ····················· ···················· ···················· ···················· |
| A | 5099 | hCoV-19/Spain/MD-H12-25/2020\|EPI_ISL_428688\|2020-0 | ···················· ···················· ····················· ···················· ···················· ···················· |
| A | 5100 | hCoV-19/Spain/MD-H12-26/2020\|EPI_ISL_428689\|2020-0 | ···················· ···················· ····················· ···················· ···················· ···················· |
| A | 5101 | hCoV-19/Spain/MD-H12-27/2020\|EPI_ISL_428690\|2020-0 | ···················· ···················· ····················· ···················· ···················· ···················· |
| A | 5102 | hCoV-19/Spain/MD-H12-28/2020\|EPI_ISL_428691\|2020-0 | ···················· ···················· ····················· ···················· ···················· ···················· |
| A | 5103 | hCoV-19/Spain/MD-H12-29/2020\|EPI_ISL_428692\|2020-0 | ···················· ···················· ····················· ···················· ···················· ···················· |
| A | 5104 | hCoV-19/Spain/MD-H12-30/2020\|EPI_ISL_428693\|2020-0 | ···················· ···················· ····················· ···················· ···················· ···················· |
| A | 5105 | hCoV-19/Spain/MD-H12-31/2020\|EPI_ISL_428694\|2020-0 | ···················· ···················· ····················· ···················· ···················· ···················· |
| A | 5106 | hCoV-19/Spain/MD-H12-32/2020\|EPI_ISL_428695\|2020-0 | ···················· ···················· ····················· ···················· ···················· ···················· |
| A | 5107 | hCoV-19/Spain/MD-H12-33/2020\|EPI_ISL_428696\|2020-0 | ···················· ···················· ····················· ···················· ···················· ···················· |
| A | 5108 | hCoV-19/Spain/MD-H12-34/2020\|EPI_ISL_428697\|2020-0 | ···················· ···················· ····················· ···················· ···················· ···················· |
| A | 5109 | hCoV-19/Spain/MD-H12-35/2020\|EPI_ISL_428698\|2020-0 | ···················· ···················· ····················· ···················· ···················· ···················· |
| A | 5110 | hCoV-19/Spain/MD-H12-36/2020\|EPI_ISL_428699\|2020-0 | ···················· ···················· ····················· ···················· ···················· ···················· |
| A | 5111 | hCoV-19/USA/CA-SR007/2020\|EPI_ISL_429990\|2020-03-2 | ···················· ···················· ····················· ···················· ···················· ···················· |
| A | 5112 | hCoV-19/USA/CA-SR017/2020\|EPI_ISL_429991\|2020-03-2 | ···················· ···················· ····················· ···················· ···················· ···················· |
| A | 5113 | hCoV-19/Jordan/SR-032/2020\|EPI_ISL_429992\|2020-03- | ···················· ···················· ····················· ···················· ···················· ···················· |
| A | 5114 | hCoV-19/Jordan/SR-033/2020\|EPI_ISL_429993\|2020-03- | ···················· ···················· ····················· ···················· ···················· ···················· |
| A | 5115 | hCoV-19/Jordan/SR-034/2020\|EPI_ISL_429994\|2020-03- | ···················· ···················· ····················· ···················· ···················· ···················· |
| A | 5117 | hCoV-19/Jordan/SR-036/2020\|EPI_ISL_429996\|2020-03- | ···················· ···················· ····················· ···················· ···················· ···················· |
| A | 5118 | hCoV-19/Jordan/SR-037/2020\|EPI_ISL_429997\|2020-03- | ···················· ·············A······ ····················· ···················· ···················· ···················· |
| A | 5119 | hCoV-19/Jordan/SR-039/2020\|EPI_ISL_429998\|2020-03- | ···················· ···················· ····················· ···················· ···················· ···················· |
| A | 5120 | hCoV-19/Jordan/SR-041/2020\|EPI_ISL_429999\|2020-03- | ···················· ···················· ····················· ···················· ···················· ···················· |
| A | 5121 | hCoV-19/France/ARA-739/2020\|EPI_ISL_410486\|2020-02 | ···················· ···················· ····················· ···················· ···················· ···················· |
| A | 5122 | hCoV-19/Jiangsu/JS01/2020\|EPI_ISL_411950\|2020-01-2 | ···················· ···················· ····················· ···················· ···················· ···················· |
| A | 5123 | hCoV-19/Sweden/01/2020\|EPI_ISL_411951\|2020-02-07 | ···················· ···················· ····················· ···················· ···················· ···················· |
| A | 5124 | hCoV-19/Jiangsu/JS02/2020\|EPI_ISL_411952\|2020-01-2 | ···················· ···················· ····················· ···················· ···················· ···················· |
| A | 5125 | hCoV-19/Jiangsu/JS03/2020\|EPI_ISL_411953\|2020-01-2 | ···················· ···················· ····················· ···················· ···················· ···················· |
| A | 5126 | hCoV-19/USA/CA-CDC-7/2020\|EPI_ISL_411954\|2020-02-0 | ···················· ···················· ····················· ···················· ···················· ···················· |
| A | 5127 | hCoV-19/USA/CA-CDC-8/2020\|EPI_ISL_411955\|2020-02-1 | ···················· ···················· ····················· ···················· ···················· ···················· |
| A | 5128 | hCoV-19/USA/TX1/2020\|EPI_ISL_411956\|2020-02-11 | ···················· ···················· ····················· ···················· ···················· ···················· |
| A | 5129 | hCoV-19/England/200940527/2020\|EPI_ISL_414005\|2020 | ···················· ···················· ····················· ···················· ···················· ···················· |
| A | 5130 | hCoV-19/England/200990724/2020\|EPI_ISL_414006\|2020 | ···················· ···················· ····················· ···················· ···················· ···················· |
| A | 5131 | hCoV-19/England/200990725/2020\|EPI_ISL_414007\|2020 | ···················· ···················· ····················· ···················· ···················· ···················· |
| A | 5132 | hCoV-19/England/200960041/2020\|EPI_ISL_414008\|2020 | ···················· ···················· ····················· ···················· ···················· ···················· |
| A | 5133 | hCoV-19/England/200960515/2020\|EPI_ISL_414009\|2020 | ···················· ···················· ····················· ···················· ···················· ···················· |
| A | 5134 | hCoV-19/England/200981386/2020\|EPI_ISL_414010\|2020 | ···T················ ···················· ····················· ···················· ···················· ···················· |
| A | 5135 | hCoV-19/England/200990006/2020\|EPI_ISL_414011\|2020 | ···················· ···················· ····················· ···················· ···················· ···················· |
| A | 5136 | hCoV-19/England/200990723/2020\|EPI_ISL_414012\|2020 | ···················· ···················· ····················· ···················· ···················· ···················· |
| A | 5137 | hCoV-19/England/201000003/2020\|EPI_ISL_414013\|2020 | ···················· ···················· ····················· ···················· ···················· ···················· |
| A | 5138 | hCoV-19/Brazil/SP-03/2020\|EPI_ISL_414014\|2020-03-0 | ···················· ···················· ····················· ···················· ···················· ···················· |
| A | 5139 | hCoV-19/Brazil/SP-06/2020\|EPI_ISL_414015\|2020-02-2 | ···················· ···················· ····················· ···················· ···················· ···················· |
| A | 5140 | hCoV-19/Brazil/SP-05/2020\|EPI_ISL_414016\|2020-02-2 | ···················· ···················· ····················· ···················· ···················· ···················· |
| A | 5141 | hCoV-19/Brazil/SP-04/2020\|EPI_ISL_414017\|2020-03-0 | ···················· ···················· ····················· ···················· ···················· ···················· |
| A | 5142 | hCoV-19/Switzerland/GE-SNRCI-29943121/2020\|EPI_ISL | ···················· ···················· ····················· ···················· ···················· ···················· |
| A | 5143 | hCoV-19/Japan/DP0699/2020\|EPI_ISL_416617\|2020-02-1 | ···················· ···················· ····················· ···················· ···················· ···················· |
| A | 5144 | hCoV-19/Japan/DP0700/2020\|EPI_ISL_416618\|2020-02-1 | ···················· ···················· ····················· ···················· ···················· ···················· |
| A | 5145 | hCoV-19/Japan/DP0703/2020\|EPI_ISL_416619\|2020-02-1 | ···················· ···················· ····················· ···················· ···················· ···················· |
| A | 5146 | hCoV-19/Japan/DP0724/2020\|EPI_ISL_416620\|2020-02-1 | ···················· ···················· ····················· ···················· ···················· ···················· |
| A | 5147 | hCoV-19/Japan/DP0743/2020\|EPI_ISL_416621\|2020-02-1 | ···················· ···················· ····················· ···················· ···················· ···················· |
| A | 5148 | hCoV-19/Japan/DP0752/2020\|EPI_ISL_416622\|2020-02-1 | ···················· ···················· ····················· ···················· ···················· ···················· |
| A | 5149 | hCoV-19/Japan/DP0764/2020\|EPI_ISL_416624\|2020-02-1 | ···················· ···················· ····················· ···················· ···················· ···················· |
| A | 5150 | hCoV-19/Japan/DP0765/2020\|EPI_ISL_416625\|2020-02-1 | ···················· ···················· ····················· ···················· ···················· ···················· |
| A | 5151 | hCoV-19/Japan/DP0779/2020\|EPI_ISL_416626\|2020-02-1 | ···················· ···················· ····················· ···················· ···················· ···················· |
| A | 5152 | hCoV-19/Japan/DP0785/2020\|EPI_ISL_416627\|2020-02-1 | ···················· ···················· ····················· ···················· ···················· ···················· |
| A | 5153 | hCoV-19/Japan/DP0786/2020\|EPI_ISL_416628\|2020-02-1 | ···················· ···················· ····················· ···················· ···················· ···················· |
| A | 5154 | hCoV-19/Japan/DP0802/2020\|EPI_ISL_416629\|2020-02-1 | ···················· ···················· ····················· ···················· ···················· ···················· |
| A | 5155 | hCoV-19/Japan/DP0890/2020\|EPI_ISL_416634\|2020-02-1 | ···················· ···················· ····················· ···················· ···················· ···················· |
| A | 5156 | hCoV-19/USA/WA-UW97/2020\|EPI_ISL_416635\|2020-03-12 | ···················· ···················· ····················· ···················· ···················· ···················· |
| A | 5157 | hCoV-19/USA/WA-UW100/2020\|EPI_ISL_416638\|2020-03-1 | ···················· ···················· ····················· ···················· ···················· ···················· |
| A | 5158 | hCoV-19/USA/WA-UW101/2020\|EPI_ISL_416639\|2020-03-1 | ···················· ···················· ····················· ···················· ···················· ···················· |
| A | 5159 | hCoV-19/USA/WA-UW103/2020\|EPI_ISL_416641\|2020-03-1 | ···················· ···················· ····················· ···················· ···················· ···················· |
| A | 5160 | hCoV-19/USA/WA-UW104/2020\|EPI_ISL_416642\|2020-03-1 | ···················· ···················· ····················· ···················· ···················· ···················· |
| A | 5161 | hCoV-19/USA/WA-UW105/2020\|EPI_ISL_416643\|2020-03-1 | ···················· ···················· ····················· ···················· ···················· ···················· |
| A | 5162 | hCoV-19/USA/WA-UW106/2020\|EPI_ISL_416644\|2020-03-1 | ···················· ···················· ····················· ···················· ···················· ···················· |
| A | 5163 | hCoV-19/USA/WA-UW108/2020\|EPI_ISL_416646\|2020-03-1 | ···················· ···················· ····················· ···················· ···················· ···················· |
| A | 5164 | hCoV-19/USA/WA-UW109/2020\|EPI_ISL_416647\|2020-03-1 | ···················· ···················· ····················· ···················· ···················· ···················· |
| A | 5165 | hCoV-19/USA/WA-UW110/2020\|EPI_ISL_416648\|2020-03-1 | ···················· ···················· ····················· ···················· ···················· ···················· |
| A | 5166 | hCoV-19/USA/WA-UW111/2020\|EPI_ISL_416649\|2020-03-1 | ···················· ···················· ····················· ···················· ···················· ···················· |
| A | 5167 | hCoV-19/USA/WA-UW112/2020\|EPI_ISL_416650\|2020-03-1 | ···················· ···················· ····················· ···················· ···················· ···················· |
| A | 5168 | hCoV-19/USA/WA-UW113/2020\|EPI_ISL_416651\|2020-03-1 | ···················· ···················· ····················· ···················· ···················· ···················· |
| A | 5169 | hCoV-19/USA/WA-UW114/2020\|EPI_ISL_416652\|2020-03-1 | ···················· ···················· ····················· ···················· ···················· ···················· |
| A | 5170 | hCoV-19/USA/WA-UW115/2020\|EPI_ISL_416653\|2020-03-1 | ···················· ···················· ····················· ···················· ···················· ···················· |
| A | 5171 | hCoV-19/USA/WA-UW116/2020\|EPI_ISL_416654\|2020-03-1 | ···················· ···················· ····················· ···················· ···················· ···················· |
| A | 5172 | hCoV-19/USA/WA-UW117/2020\|EPI_ISL_416655\|2020-03-1 | ···················· ···················· ····················· ···················· ···················· ···················· |
| A | 5173 | hCoV-19/USA/WA-UW118/2020\|EPI_ISL_416656\|2020-03-1 | ···················· ···················· ····················· ···················· ···················· ···················· |

| | | | | | | | | |
|---|---|---|---|---|---|---|---|---|
| A | 5174 | hCoV-19/USA/WA-UW119/2020\|EPI_ISL_416657\|2020-03-1 | .................... | .................... | ..................... | .................... | .................... | .................... |
| A | 5175 | hCoV-19/USA/WA-UW120/2020\|EPI_ISL_416658\|2020-03-1 | .................... | .................... | ..................... | .................... | .................... | .................... |
| A | 5176 | hCoV-19/USA/WA-UW121/2020\|EPI_ISL_416659\|2020-03-1 | .................... | .................... | ..................... | .................... | .................... | .................... |
| A | 5177 | hCoV-19/USA/WA-UW122/2020\|EPI_ISL_416660\|2020-03-1 | .................... | .................... | ..................... | .................... | .................... | .................... |
| A | 5178 | hCoV-19/USA/WA-UW123/2020\|EPI_ISL_416661\|2020-03-1 | .................... | .................... | ..................... | .................... | .................... | .................... |
| A | 5179 | hCoV-19/USA/WA-UW124/2020\|EPI_ISL_416662\|2020-03-1 | .................... | .................... | ..................... | .................... | .................... | .................... |
| A | 5180 | hCoV-19/USA/WA-UW125/2020\|EPI_ISL_416663\|2020-03-1 | .................... | .................... | ..................... | .................... | .................... | .................... |
| A | 5181 | hCoV-19/USA/WA-UW126/2020\|EPI_ISL_416664\|2020-03-1 | .................... | .................... | ..................... | .................... | .................... | .................... |
| A | 5182 | hCoV-19/USA/WA-UW127/2020\|EPI_ISL_416665\|2020-03-1 | .................... | .................... | ..................... | .................... | .................... | .................... |
| A | 5183 | hCoV-19/USA/WA-UW128/2020\|EPI_ISL_416666\|2020-03-1 | .................... | .................... | ..................... | .................... | .................... | .................... |
| A | 5184 | hCoV-19/USA/WA-UW129/2020\|EPI_ISL_416667\|2020-03-1 | .................... | .................... | ..................... | .................... | .................... | .................... |
| A | 5185 | hCoV-19/USA/WA-UW130/2020\|EPI_ISL_416668\|2020-03-1 | .................... | .................... | ..................... | .................... | .................... | .................... |
| A | 5186 | hCoV-19/USA/WA-UW131/2020\|EPI_ISL_416669\|2020-03-1 | .................... | .................... | ..................... | .................... | .................... | .................... |
| A | 5187 | hCoV-19/USA/WA-UW133/2020\|EPI_ISL_416671\|2020-03-1 | .................... | .................... | ..................... | .................... | .................... | .................... |
| A | 5188 | hCoV-19/USA/OR-UW135/2020\|EPI_ISL_416673\|2020-03-1 | .................... | .................... | ..................... | .................... | .................... | .................... |
| A | 5189 | hCoV-19/USA/WA-UW136/2020\|EPI_ISL_416674\|2020-03-1 | .................... | .................... | ..................... | .................... | .................... | .................... |
| A | 5190 | hCoV-19/USA/WA-UW137/2020\|EPI_ISL_416675\|2020-03-1 | .................... | .................... | ..................... | .................... | .................... | .................... |
| A | 5191 | hCoV-19/USA/WA-UW138/2020\|EPI_ISL_416676\|2020-03-1 | .................... | .................... | ..................... | .................... | .................... | .................... |
| A | 5192 | hCoV-19/USA/WA-UW139/2020\|EPI_ISL_416677\|2020-03-1 | .................... | .................... | ..................... | .................... | .................... | .................... |
| A | 5193 | hCoV-19/USA/WA-UW140/2020\|EPI_ISL_416678\|2020-03-1 | .................... | .................... | ..................... | .................... | .................... | .................... |
| A | 5194 | hCoV-19/USA/WA-UW141/2020\|EPI_ISL_416679\|2020-03-1 | .................... | .................... | ..................... | .................... | .................... | .................... |
| A | 5195 | hCoV-19/Malaysia/189332/2020\|EPI_ISL_417917\|2020-0 | .................... | .................... | ..................... | .................... | .................... | .................... |
| A | 5196 | hCoV-19/Malaysia/188407/2020\|EPI_ISL_417918\|2020-0 | .................... | .................... | ..................... | .................... | .................... | .................... |
| A | 5197 | hCoV-19/Italy/LAZ-INMI3/2020\|EPI_ISL_417921\|2020-0 | .................... | .................... | ..................... | .................... | .................... | .................... |
| A | 5198 | hCoV-19/Italy/LAZ-INMI4/2020\|EPI_ISL_417922\|2020-0 | .................... | .................... | ..................... | .................... | .................... | .................... |
| A | 5199 | hCoV-19/Italy/LAZ-INMI5/2020\|EPI_ISL_417923\|2020-0 | .................... | .................... | ..................... | .................... | .................... | .................... |
| A | 5200 | hCoV-19/Colombia/ANT-INS-79256/2020\|EPI_ISL_417924 | .................... | .................... | ..................... | .................... | .................... | .................... |
| A | 5201 | hCoV-19/DRC/94/2020\|EPI_ISL_417947\|2020-03-19 | .................... | .................... | ..................... | .................... | .................... | .................... |
| A | 5202 | hCoV-19/DRC/108/2020\|EPI_ISL_417948\|2020-03-19 | .................... | .................... | ..................... | .................... | .................... | .................... |
| A | 5203 | hCoV-19/DRC/158/2020\|EPI_ISL_417950\|2020-03-20 | .................... | .................... | ..................... | .................... | .................... | .................... |
| A | 5204 | hCoV-19/Spain/MD-HLP-H3-10/2020\|EPI_ISL_417954\|202 | .................... | .................... | ..................... | .................... | .................... | .................... |
| A | 5205 | hCoV-19/DRC/191/2020\|EPI_ISL_417955\|2020-03-21 | .................... | .................... | ..................... | .................... | .................... | .................... |
| A | 5206 | hCoV-19/USA/UT-010/2020\|EPI_ISL_417958\|2020-03-10 | .................... | .................... | ..................... | .................... | .................... | .................... |
| A | 5208 | hCoV-19/USA/UT-016/2020\|EPI_ISL_417966\|2020-03-13 | .................... | .................... | ..................... | .................... | .................... | .................... |
| A | 5209 | hCoV-19/Spain/MD-HLP-LP10-12/2020\|EPI_ISL_417969\|2 | .................... | .................... | ..................... | .................... | .................... | .................... |
| A | 5210 | hCoV-19/USA/UT-027/2020\|EPI_ISL_417970\|2020-03-16 | .................... | .................... | ..................... | .................... | .................... | .................... |
| A | 5211 | hCoV-19/USA/UT-028/2020\|EPI_ISL_417971\|2020-03-19 | .................... | .................... | ..................... | .................... | .................... | .................... |
| A | 5212 | hCoV-19/Spain/MD-HLP-LP12-21/2020\|EPI_ISL_417972\|2 | .................... | .................... | ..................... | .................... | .................... | .................... |
| A | 5213 | hCoV-19/USA/UT-031/2020\|EPI_ISL_417973\|2020-03-19 | .................... | .................... | ..................... | .................... | .................... | .................... |
| A | 5214 | hCoV-19/USA/UT-032/2020\|EPI_ISL_417974\|2020-03-19 | .................... | .................... | ..................... | .................... | .................... | .................... |
| A | 5215 | hCoV-19/Spain/MD-HLP-LP14-3/2020\|EPI_ISL_417975\|20 | .................... | .................... | ..................... | .................... | .................... | .................... |
| A | 5216 | hCoV-19/USA/UT-033/2020\|EPI_ISL_417976\|2020-03-19 | .................... | .................... | ..................... | .................... | .................... | .................... |
| A | 5217 | hCoV-19/USA/UT-034/2020\|EPI_ISL_417977\|2020-03-20 | .................... | .................... | ..................... | .................... | .................... | .................... |
| A | 5218 | hCoV-19/Spain/MD-HLP-LP15-4/2020\|EPI_ISL_417978\|20 | .................... | .................... | ..................... | .................... | .................... | .................... |
| A | 5219 | hCoV-19/Spain/MD-HLP-R2-15/2020\|EPI_ISL_417979\|202 | .................... | .................... | ..................... | .................... | .................... | .................... |
| A | 5220 | hCoV-19/Spain/MD-HLP-R5-8/2020\|EPI_ISL_417980\|2020 | .................... | .................... | ..................... | .................... | .................... | .................... |
| A | 5221 | hCoV-19/Spain/MD-HLP-R10-33/2020\|EPI_ISL_417981\|20 | .................... | .................... | ..................... | .................... | .................... | .................... |
| A | 5222 | hCoV-19/Portugal/PT0001b/2020\|EPI_ISL_417986\|2020- | .................... | .................... | ..................... | .................... | .................... | .................... |
| A | 5223 | hCoV-19/Portugal/PT0003/2020\|EPI_ISL_417987\|2020-0 | .................... | .................... | ..................... | .................... | .................... | .................... |
| A | 5224 | hCoV-19/Portugal/PT0004/2020\|EPI_ISL_417988\|2020-0 | .................... | .................... | ..................... | .................... | .................... | .................... |
| A | 5225 | hCoV-19/Portugal/PT0005/2020\|EPI_ISL_417989\|2020-0 | .................... | .................... | ..................... | .................... | .................... | .................... |
| A | 5226 | hCoV-19/Portugal/PT0006a/2020\|EPI_ISL_417990\|2020- | .................... | .................... | ..................... | .................... | .................... | .................... |
| A | 5227 | hCoV-19/Portugal/PT0006b/2020\|EPI_ISL_417991\|2020- | .................... | .................... | ..................... | .................... | .................... | .................... |
| A | 5228 | hCoV-19/Portugal/PT0007/2020\|EPI_ISL_417992\|2020-0 | .................... | .................... | ..................... | .................... | .................... | .................... |
| A | 5229 | hCoV-19/Portugal/PT0008/2020\|EPI_ISL_417993\|2020-0 | .................... | .................... | ..................... | .................... | .................... | .................... |
| A | 5230 | hCoV-19/Portugal/PT0009/2020\|EPI_ISL_417994\|2020-0 | .................... | .................... | ..................... | .................... | .................... | .................... |
| A | 5231 | hCoV-19/Portugal/PT0010/2020\|EPI_ISL_417995\|2020-0 | .................... | .................... | ..................... | .................... | .................... | .................... |
| A | 5232 | hCoV-19/Portugal/PT0011/2020\|EPI_ISL_417996\|2020-0 | .................... | .................... | ..................... | .................... | .................... | .................... |
| A | 5233 | hCoV-19/Portugal/PT0012/2020\|EPI_ISL_417997\|2020-0 | .................... | .................... | ..................... | .................... | .................... | .................... |
| A | 5234 | hCoV-19/Portugal/PT0013/2020\|EPI_ISL_417998\|2020-0 | .................... | .................... | ..................... | .................... | .................... | .................... |
| A | 5235 | hCoV-19/Portugal/PT0014/2020\|EPI_ISL_417999\|2020-0 | .................... | .................... | ..................... | .................... | .................... | .................... |
| A | 5236 | hCoV-19/Spain/MD-H12-1301/2020\|EPI_ISL_421171\|2020 | .................... | .................... | ..................... | .................... | .................... | .................... |
| A | 5237 | hCoV-19/Spain/MD-H12-H12-1502/2020\|EPI_ISL_421172\| | .................... | .................... | ..................... | .................... | .................... | .................... |
| A | 5238 | hCoV-19/Spain/MD-H12-H12-1703/2020\|EPI_ISL_421173\| | .................... | .................... | ..................... | .................... | .................... | .................... |
| A | 5239 | hCoV-19/Spain/MD-H12-1804/2020\|EPI_ISL_421174\|2020 | .................... | .................... | ..................... | .................... | .................... | .................... |
| A | 5240 | hCoV-19/Spain/MD-H12-H12-1905/2020\|EPI_ISL_421175\| | .................... | .................... | ..................... | .................... | .................... | .................... |
| A | 5241 | hCoV-19/Spain/MD-H12-2006/2020\|EPI_ISL_421176\|2020 | .................... | .................... | ..................... | .................... | .................... | .................... |
| A | 5242 | hCoV-19/Spain/MD-H12-H12-2107/2020\|EPI_ISL_421177\| | .................... | .................... | ..................... | .................... | .................... | .................... |
| A | 5243 | hCoV-19/Spain/MD-H12-H12-2208/2020\|EPI_ISL_421178\| | .................... | .................... | ..................... | .................... | .................... | .................... |
| A | 5244 | hCoV-19/Spain/MD-H12-H12-2309/2020\|EPI_ISL_421179\| | .................... | .................... | ..................... | .................... | .................... | .................... |
| A | 5245 | hCoV-19/Spain/MD-H12-2410/2020\|EPI_ISL_421180\|2020 | .................... | .................... | ..................... | .................... | .................... | .................... |
| A | 5246 | hCoV-19/Belgium/ULG-10001/2020\|EPI_ISL_421182\|2020 | .................... | .................... | ..................... | .................... | .................... | .................... |
| A | 5247 | hCoV-19/Belgium/ULG-10003/2020\|EPI_ISL_421183\|2020 | .................... | .................... | ..................... | .................... | .................... | .................... |
| A | 5248 | hCoV-19/Belgium/ULG-10004/2020\|EPI_ISL_421184\|2020 | .................... | .................... | ..................... | .................... | .................... | .................... |
| A | 5249 | hCoV-19/Belgium/ULG-10006/2020\|EPI_ISL_421185\|2020 | .................... | .................... | ..................... | .................... | .................... | .................... |
| A | 5250 | hCoV-19/Belgium/ULG-10007/2020\|EPI_ISL_421186\|2020 | .................... | .................... | ..................... | .................... | .................... | .................... |
| A | 5251 | hCoV-19/Belgium/ULG-10008/2020\|EPI_ISL_421187\|2020 | .................... | .................... | ..................... | .................... | .................... | .................... |
| A | 5252 | hCoV-19/Belgium/ULG-10009/2020\|EPI_ISL_421188\|2020 | .................... | .................... | ..................... | .................... | .................... | .................... |

| | | | | | | | |
|---|---|---|---|---|---|---|---|
| A | 5253 hCoV-19/Belgium/ULG-10011/2020\|EPI_ISL_421189\|2020 | ···················· | ···················· | ···················· | ···················· | ···················· | ···················· |
| A | 5254 hCoV-19/USA/WI-UW-42/2020\|EPI_ISL_421300\|2020-03-1 | ···················· | ···················· | ···················· | ···················· | ···················· | ···················· |
| A | 5255 hCoV-19/USA/WI-UW-43/2020\|EPI_ISL_421301\|2020-03-1 | ···················· | ···················· | ···················· | ···················· | ···················· | ···················· |
| A | 5256 hCoV-19/Singapore/42/2020\|EPI_ISL_422435\|2020-03-3 | ···················· | ···················· | ···················· | ···················· | ···················· | ···················· |
| A | 5257 hCoV-19/Italy/VEN-IZSVe-21-26/2020\|EPI_ISL_422437\| | ···················· | ···················· | ···················· | ···················· | ···················· | ···················· |
| A | 5258 hCoV-19/Italy/VEN-IZSVe-21-37/2020\|EPI_ISL_422438\| | ···················· | ···················· | ···················· | ···················· | ···················· | ···················· |
| A | 5259 hCoV-19/USA/WI-GMF-00498/2020\|EPI_ISL_422453\|2020- | ···················· | ···················· | ···················· | ···················· | ···················· | ···················· |
| A | 5260 hCoV-19/USA/WI-GMF-00557/2020\|EPI_ISL_422459\|2020- | ···················· | ···················· | ···················· | ···················· | ···················· | ···················· |
| A | 5261 hCoV-19/USA/WI-GMF-00707/2020\|EPI_ISL_422461\|2020- | ···················· | ···················· | ···················· | ···················· | ···················· | ···················· |
| A | 5262 hCoV-19/USA/WI-GMF-00744/2020\|EPI_ISL_422462\|2020- | ···················· | ···················· | ···················· | ···················· | ···················· | ···················· |
| A | 5263 hCoV-19/USA/WI-GMF-00588/2020\|EPI_ISL_422463\|2020- | ···················· | ···················· | ···················· | ···················· | ···················· | ···················· |
| A | 5264 hCoV-19/USA/WI-GMF-00441/2020\|EPI_ISL_422465\|2020- | ···················· | ···················· | ···················· | ···················· | ···················· | ···················· |
| A | 5265 hCoV-19/USA/NY-PV09045/2020\|EPI_ISL_422488\|2020-03 | ···················· | ···················· | ···················· | ···················· | ···················· | ···················· |
| A | 5266 hCoV-19/USA/NY-PV09057/2020\|EPI_ISL_422489\|2020-03 | ···················· | ···················· | ···················· | ···················· | ···················· | ···················· |
| A | 5267 hCoV-19/USA/NY-PV09069/2020\|EPI_ISL_422490\|2020-03 | ···················· | ···················· | ···················· | ···················· | ···················· | ···················· |
| A | 5268 hCoV-19/USA/NY-PV09115/2020\|EPI_ISL_422491\|2020-03 | ···················· | ···················· | ···················· | ···················· | ···················· | ···················· |
| A | 5269 hCoV-19/USA/NY-PV09116/2020\|EPI_ISL_422492\|2020-03 | ···················· | ···················· | ···················· | ···················· | ···················· | ···················· |
| A | 5270 hCoV-19/USA/NY-PV09119/2020\|EPI_ISL_422494\|2020-03 | ···················· | ···················· | ···················· | ···················· | ···················· | ···················· |
| A | 5271 hCoV-19/USA/NY-PV09120/2020\|EPI_ISL_422495\|2020-03 | ···················· | ···················· | ···················· | ···················· | ···················· | ···················· |
| A | 5272 hCoV-19/USA/NY-PV09121/2020\|EPI_ISL_422496\|2020-03 | ···················· | ···················· | ···················· | ···················· | ···················· | ···················· |
| A | 5273 hCoV-19/USA/NY-PV09122/2020\|EPI_ISL_422497\|2020-03 | ···················· | ···················· | ···················· | ···················· | ···················· | ···················· |
| A | 5274 hCoV-19/USA/NY-PV09123/2020\|EPI_ISL_422498\|2020-03 | ···················· | ···················· | ···················· | ···················· | ···················· | ···················· |
| A | 5275 hCoV-19/USA/NY-PV09125/2020\|EPI_ISL_422499\|2020-03 | ···················· | ···················· | ···················· | ···················· | ···················· | ···················· |
| A | 5276 hCoV-19/Netherlands/NA_291/2020\|EPI_ISL_422600\|202 | ···················· | ···················· | ···················· | ···················· | ···················· | ···················· |
| A | 5277 hCoV-19/Netherlands/NA_292/2020\|EPI_ISL_422601\|202 | ···················· | ···················· | ···················· | ···················· | ···················· | ···················· |
| A | 5278 hCoV-19/Netherlands/NA_293/2020\|EPI_ISL_422602\|202 | ···················· | ···················· | ···················· | ···················· | ···················· | ···················· |
| A | 5279 hCoV-19/Netherlands/NA_294/2020\|EPI_ISL_422603\|202 | ···················· | ···················· | ···················· | ···················· | ···················· | ···················· |
| A | 5280 hCoV-19/Netherlands/NA_295/2020\|EPI_ISL_422604\|202 | ···················· | ···················· | ···················· | ···················· | ···················· | ···················· |
| A | 5281 hCoV-19/Netherlands/NA_296/2020\|EPI_ISL_422605\|202 | ···················· | ···················· | ···················· | ···················· | ···················· | ···················· |
| A | 5282 hCoV-19/Netherlands/NA_297/2020\|EPI_ISL_422606\|202 | ···················· | ···················· | ···················· | ···················· | ···················· | ···················· |
| A | 5283 hCoV-19/Netherlands/NA_298/2020\|EPI_ISL_422607\|202 | ···················· | ···················· | ···················· | ···················· | ···················· | ···················· |
| A | 5284 hCoV-19/Netherlands/NA_299/2020\|EPI_ISL_422608\|202 | ···················· | ···················· | ···················· | ···················· | ···················· | ···················· |
| A | 5285 hCoV-19/Netherlands/NA_300/2020\|EPI_ISL_422609\|202 | ···················· | ···················· | ···················· | ···················· | ···················· | ···················· |
| A | 5286 hCoV-19/Netherlands/NA_301/2020\|EPI_ISL_422610\|202 | ···················· | ···················· | ···················· | ···················· | ···················· | ···················· |
| A | 5287 hCoV-19/Netherlands/NA_313/2020\|EPI_ISL_422622\|202 | ···················· | ···················· | ···················· | ···················· | ···················· | ···················· |
| A | 5288 hCoV-19/Netherlands/NA_316/2020\|EPI_ISL_422625\|202 | ···················· | ···················· | ···················· | ···················· | ···················· | ···················· |
| A | 5289 hCoV-19/Netherlands/NA_317/2020\|EPI_ISL_422626\|202 | ···················· | ···················· | ···················· | ···················· | ···················· | ···················· |
| A | 5290 hCoV-19/Netherlands/ZuidHolland_100/2020\|EPI_ISL_4 | ···················· | ···········A········ | ···················· | ···················· | ···················· | ···················· |
| A | 5291 hCoV-19/Netherlands/ZuidHolland_101/2020\|EPI_ISL_4 | ···················· | ···················· | ···················· | ···················· | ···················· | ···················· |
| A | 5292 hCoV-19/Netherlands/ZuidHolland_94/2020\|EPI_ISL_42 | ···················· | ···················· | ···················· | ···················· | ···················· | ···················· |
| A | 5293 hCoV-19/Netherlands/ZuidHolland_95/2020\|EPI_ISL_42 | ···················· | ···················· | ···················· | ···················· | ···················· | ···················· |
| A | 5294 hCoV-19/Netherlands/ZuidHolland_96/2020\|EPI_ISL_42 | ···················· | ···················· | ···················· | ···················· | ···················· | ···················· |
| A | 5295 hCoV-19/England/20132031404/2020\|EPI_ISL_423769\|20 | ···················· | ···················· | ···················· | ···················· | ···················· | ···················· |
| A | 5296 hCoV-19/England/20132032504/2020\|EPI_ISL_423771\|20 | ···················· | ···················· | ···················· | ···················· | ···················· | ···················· |
| A | 5297 hCoV-19/England/20132032604/2020\|EPI_ISL_423772\|20 | ···················· | ···················· | ···················· | ···················· | ···················· | ···················· |
| A | 5299 hCoV-19/England/20132032804/2020\|EPI_ISL_423774\|20 | ···················· | ···················· | ···················· | ···················· | ···················· | ···················· |
| A | 5300 hCoV-19/England/20132032904/2020\|EPI_ISL_423775\|20 | ···················· | ···················· | ···················· | ···················· | ···················· | ···················· |
| A | 5302 hCoV-19/England/20132035904/2020\|EPI_ISL_423779\|20 | ···················· | ···················· | ···················· | ···················· | ···················· | ···················· |
| A | 5303 hCoV-19/England/20132036004/2020\|EPI_ISL_423780\|20 | ···················· | ···················· | ···················· | ···················· | ···················· | ···················· |
| A | 5304 hCoV-19/England/20132047304/2020\|EPI_ISL_423782\|20 | ···················· | ···················· | ···················· | ···················· | ···················· | ···················· |
| A | 5306 hCoV-19/England/20132054604/2020\|EPI_ISL_423786\|20 | ···················· | ···················· | ···················· | ···················· | ···················· | ···················· |
| A | 5307 hCoV-19/England/20132054704/2020\|EPI_ISL_423787\|20 | ···················· | ···················· | ···················· | ···················· | ···················· | ···················· |
| A | 5308 hCoV-19/England/20132066404/2020\|EPI_ISL_423790\|20 | ···················· | ···················· | ···················· | ···················· | ···················· | ···················· |
| A | 5310 hCoV-19/England/20132068204/2020\|EPI_ISL_423796\|20 | ···················· | ···················· | ···················· | ···················· | ···················· | ···················· |
| A | 5311 hCoV-19/England/20132068304/2020\|EPI_ISL_423797\|20 | ···················· | ···················· | ···················· | ···················· | ···················· | ···················· |
| A | 5312 hCoV-19/England/20132069004/2020\|EPI_ISL_423798\|20 | ···················· | ···················· | ···················· | ···················· | ···················· | ··N···········N····· |
| A | 5313 hCoV-19/England/20132069104/2020\|EPI_ISL_423799\|20 | ···················· | ···················· | ···················· | ···················· | ···················· | ···················· |
| A | 5314 hCoV-19/England/20128002204/2020\|EPI_ISL_423923\|20 | ···················· | ···················· | ···················· | ···················· | ···················· | ···················· |
| A | 5315 hCoV-19/England/20128003004/2020\|EPI_ISL_423924\|20 | ···················· | ···················· | ···················· | ···················· | ···················· | ···················· |
| A | 5316 hCoV-19/England/20128003506/2020\|EPI_ISL_423926\|20 | ···················· | ···················· | ···················· | ···················· | ···················· | ···················· |
| A | 5317 hCoV-19/England/20128003904/2020\|EPI_ISL_423927\|20 | ···················· | ···················· | ···················· | ···················· | ···················· | ···················· |
| A | 5319 hCoV-19/England/20128011804/2020\|EPI_ISL_423933\|20 | ···················· | ···················· | ···················· | ···················· | ···················· | ···················· |
| A | 5320 hCoV-19/England/20128012704/2020\|EPI_ISL_423935\|20 | ···················· | ···················· | ···················· | ···················· | ···················· | ···················· |
| A | 5325 hCoV-19/England/20128041604/2020\|EPI_ISL_423963\|20 | ···················· | ···················· | ···················· | ···················· | ···················· | ···················· |
| A | 5326 hCoV-19/Brazil/RJ-477/2020\|EPI_ISL_427300\|2020-03- | ···················· | ···················· | ···················· | ···················· | ···················· | ···················· |
| A | 5327 hCoV-19/Brazil/RJ-477i/2020\|EPI_ISL_427301\|2020-03 | ···················· | ···················· | ···················· | ···················· | ···················· | ···················· |
| A | 5328 hCoV-19/Brazil/RJ-763/2020\|EPI_ISL_427302\|2020-03- | ···················· | ···················· | ···················· | ···················· | ···················· | ···················· |
| A | 5329 hCoV-19/Brazil/RJ-818/2020\|EPI_ISL_427303\|2020-03- | ···················· | ···················· | ···················· | ···················· | ···················· | ···················· |
| A | 5330 hCoV-19/Brazil/RJ-872/2020\|EPI_ISL_427304\|2020-03- | ···················· | ···················· | ···················· | ···················· | ···················· | ···················· |
| A | 5331 hCoV-19/Brazil/SC-766/2020\|EPI_ISL_427305\|2020-03- | ···················· | ···················· | ···················· | ···················· | ···················· | ···················· |
| A | 5332 hCoV-19/Brazil/SC-769/2020\|EPI_ISL_427306\|2020-03- | ···················· | ···················· | ···················· | ···················· | ···················· | ···················· |
| A | 5333 hCoV-19/Russia/StPetersburg-RII4144S/2020\|EPI_ISL_ | ···················· | ···················· | ···················· | ···················· | ···················· | ···················· |
| A | 5334 hCoV-19/Russia/StPetersburg-RII4307V/2020\|EPI_ISL_ | ···················· | ···················· | ···················· | ···················· | ···················· | ···················· |
| A | 5335 hCoV-19/Russia/StPetersburg-RII4308V/2020\|EPI_ISL_ | ···················· | ···················· | ···················· | ···················· | ···················· | ···················· |
| A | 5336 hCoV-19/Russia/StPetersburg-RII4382V/2020\|EPI_ISL_ | ···················· | ···················· | ···················· | ···················· | ···················· | ···················· |
| A | 5337 hCoV-19/Russia/StPetersburg-RII4384V/2020\|EPI_ISL_ | ···················· | ···················· | ···················· | ···················· | ···················· | ···················· |
| A | 5338 hCoV-19/Russia/StPetersburg-RII4386V/2020\|EPI_ISL_ | ···················· | ···················· | ···················· | ···················· | ···················· | ···················· |
| A | 5339 hCoV-19/Russia/StPetersburg-RII4532V/2020\|EPI_ISL_ | ···················· | ···················· | ···················· | ···················· | ···················· | ···················· |

| | | | | | | | | |
|---|---|---|---|---|---|---|---|---|
| A | 5340 | hCoV-19/Russia/StPetersburg-RII4546V/2020\|EPI_ISL_ | .................... | .................... | ..................... | .................... | .................... | .................... |
| A | 5341 | hCoV-19/Russia/Ulan-Ude-RII4560S/2020\|EPI_ISL_4273 | .................... | .................... | ..................... | .................... | .................... | .................... |
| A | 5342 | hCoV-19/Russia/StPetersburg-RII4584S/2020\|EPI_ISL_ | .................... | .................... | ..................... | .................... | .................... | .................... |
| A | 5343 | hCoV-19/Russia/StPetersburg-RII4585S/2020\|EPI_ISL_ | .................... | .................... | ..................... | .................... | .................... | .................... |
| A | 5344 | hCoV-19/Russia/StPetersburg-RII4591S/2020\|EPI_ISL_ | .................... | .................... | ..................... | .................... | .................... | .................... |
| A | 5345 | hCoV-19/Russia/StPetersburg-RII4597S/2020\|EPI_ISL_ | .................... | .................... | ..................... | .................... | .................... | .................... |
| A | 5346 | hCoV-19/Russia/StPetersburg-RII4637S/2020\|EPI_ISL_ | .................... | .................... | ..................... | .................... | .................... | .................... |
| A | 5347 | hCoV-19/Russia/StPetersburg-RII4649S/2020\|EPI_ISL_ | .................... | .................... | ..................... | .................... | .................... | .................... |
| A | 5348 | hCoV-19/Russia/StPetersburg-RII4655S/2020\|EPI_ISL_ | .................... | .................... | ..................... | .................... | .................... | .................... |
| A | 5349 | hCoV-19/Russia/StPetersburg-RII4678S/2020\|EPI_ISL_ | .................... | .................... | ..................... | .................... | .................... | .................... |
| A | 5350 | hCoV-19/Russia/StPetersburg-RII4686S/2020\|EPI_ISL_ | .................... | .................... | ..................... | .................... | .................... | .................... |
| A | 5351 | hCoV-19/Russia/StPetersburg-RII4693S/2020\|EPI_ISL_ | .................... | .................... | ..................... | .................... | .................... | .................... |
| A | 5352 | hCoV-19/Russia/StPetersburg-RII4694S/2020\|EPI_ISL_ | .................... | .................... | ..................... | .................... | .................... | .................... |
| A | 5353 | hCoV-19/Russia/StPetersburg-RII4696S/2020\|EPI_ISL_ | .................... | .................... | ..................... | .................... | .................... | .................... |
| A | 5354 | hCoV-19/Russia/StPetersburg-RII4697S/2020\|EPI_ISL_ | .................... | .................... | ..................... | .................... | .................... | .................... |
| A | 5355 | hCoV-19/Russia/StPetersburg-RII4707S/2020\|EPI_ISL_ | .................... | .................... | ..................... | .................... | .................... | .................... |
| A | 5356 | hCoV-19/France/HDF-1463/2020\|EPI_ISL_429968\|2020-0 | .................... | .................... | ..................... | .................... | .................... | .................... |
| A | 5357 | hCoV-19/USA/VA-DCLS-0069/2020\|EPI_ISL_429969\|2020- | .................... | .................... | ..................... | .................... | .................... | .................... |
| A | 5358 | hCoV-19/USA/VA-DCLS-0070/2020\|EPI_ISL_429970\|2020- | .................... | .................... | ..................... | .................... | .................... | .................... |
| A | 5359 | hCoV-19/USA/VA-DCLS-0071/2020\|EPI_ISL_429971\|2020- | .................... | .................... | ..................... | .................... | .................... | .................... |
| A | 5360 | hCoV-19/USA/VA-DCLS-0072/2020\|EPI_ISL_429972\|2020- | .................... | .................... | ..................... | .................... | .................... | .................... |
| A | 5361 | hCoV-19/USA/VA-DCLS-0073/2020\|EPI_ISL_429973\|2020- | .................... | .................... | ..................... | .................... | .................... | .................... |
| A | 5362 | hCoV-19/USA/VA-DCLS-0075/2020\|EPI_ISL_429974\|2020- | .................... | .................... | ..................... | .................... | .................... | .................... |
| A | 5363 | hCoV-19/USA/VA-DCLS-0076/2020\|EPI_ISL_429975\|2020- | .................... | .................... | ..................... | .................... | .................... | .................... |
| A | 5364 | hCoV-19/USA/VA-DCLS-0077/2020\|EPI_ISL_429976\|2020- | .................... | .................... | ..................... | .................... | .................... | .................... |
| A | 5365 | hCoV-19/USA/VA-DCLS-0078/2020\|EPI_ISL_429977\|2020- | .................... | .................... | ..................... | .................... | .................... | .................... |
| A | 5366 | hCoV-19/USA/VA-DCLS-0079/2020\|EPI_ISL_429978\|2020- | .................... | .................... | ..................... | .................... | .................... | .................... |
| A | 5367 | hCoV-19/USA/VA-DCLS-0080/2020\|EPI_ISL_429979\|2020- | .................... | .................... | ..................... | .................... | .................... | .................... |
| A | 5368 | hCoV-19/USA/VA-DCLS-0081/2020\|EPI_ISL_429980\|2020- | .................... | .................... | ..................... | .................... | .................... | .................... |
| A | 5369 | hCoV-19/USA/VA-DCLS-0084/2020\|EPI_ISL_429982\|2020- | .................... | .................... | ..................... | .................... | .................... | .................... |
| A | 5370 | hCoV-19/USA/VA-DCLS-0086/2020\|EPI_ISL_429983\|2020- | .................... | .................... | ..................... | .................... | .................... | .................... |
| A | 5371 | hCoV-19/USA/VA-DCLS-0087/2020\|EPI_ISL_429984\|2020- | .................... | .................... | ..................... | .................... | .................... | .................... |
| A | 5372 | hCoV-19/USA/VA-DCLS-0088/2020\|EPI_ISL_429985\|2020- | .................... | .................... | ..................... | .................... | .................... | .................... |
| A | 5373 | hCoV-19/USA/VA-DCLS-0089/2020\|EPI_ISL_429986\|2020- | .................... | .................... | ..................... | .................... | .................... | .................... |
| A | 5374 | hCoV-19/USA/VA-DCLS-0090/2020\|EPI_ISL_429987\|2020- | .................... | .................... | ..................... | .................... | .................... | .................... |
| A | 5375 | hCoV-19/USA/VA-DCLS-0091/2020\|EPI_ISL_429988\|2020- | .................... | .................... | ..................... | .................... | .................... | .................... |
| A | 5376 | hCoV-19/USA/VA-DCLS-0092/2020\|EPI_ISL_429989\|2020- | .................... | .................... | ..................... | .................... | .................... | .................... |
| A | 5377 | hCoV-19/Cambodia/0012/2020\|EPI_ISL_411902\|2020-01- | .................... | .................... | ..................... | .................... | .................... | .................... |
| A | 5378 | hCoV-19/Taiwan/CGMH-CGU-01/2020\|EPI_ISL_411915\|202 | .................... | .................... | ..................... | .................... | .................... | .................... |
| A | 5379 | hCoV-19/Taiwan/3/2020\|EPI_ISL_411926\|2020-01-24 | .................... | .................... | ..................... | .................... | .................... | .................... |
| A | 5380 | hCoV-19/Taiwan/4/2020\|EPI_ISL_411927\|2020-01-28 | .................... | .................... | ..................... | .................... | .................... | .................... |
| A | 5381 | hCoV-19/Netherlands/NA_14/2020\|EPI_ISL_415470\|2020 | .................... | .................... | ..................... | .................... | .................... | .................... |
| A | 5382 | hCoV-19/Netherlands/NA_15/2020\|EPI_ISL_415471\|2020 | .................... | .................... | ..................... | .................... | .................... | .................... |
| A | 5383 | hCoV-19/Netherlands/NA_16/2020\|EPI_ISL_415472\|2020 | .................... | .................... | ..................... | .................... | .................... | .................... |
| A | 5384 | hCoV-19/Netherlands/NA_17/2020\|EPI_ISL_415473\|2020 | .................... | .................... | ..................... | .................... | .................... | .................... |
| A | 5385 | hCoV-19/Netherlands/NA_18/2020\|EPI_ISL_415474\|2020 | .................... | .................... | ..................... | .................... | .................... | .................... |
| A | 5386 | hCoV-19/Netherlands/NA_19/2020\|EPI_ISL_415475\|2020 | .................... | .................... | ..................... | .................... | .................... | .................... |
| A | 5387 | hCoV-19/Netherlands/NA_2/2020\|EPI_ISL_415476\|2020- | .................... | .................... | ..................... | .................... | .................... | .................... |
| A | 5388 | hCoV-19/Netherlands/NA_21/2020\|EPI_ISL_415478\|2020 | .................... | .................... | ..................... | .................... | .................... | .................... |
| A | 5389 | hCoV-19/Netherlands/NA_22/2020\|EPI_ISL_415479\|2020 | .................... | .................... | ..................... | .................... | .................... | .................... |
| A | 5390 | hCoV-19/Netherlands/NA_23/2020\|EPI_ISL_415480\|2020 | .................... | .................... | ..................... | .................... | .................... | .................... |
| A | 5391 | hCoV-19/Netherlands/NA_24/2020\|EPI_ISL_415481\|2020 | .................... | .................... | ..................... | .................... | .................... | .................... |
| A | 5392 | hCoV-19/Netherlands/NA_25/2020\|EPI_ISL_415482\|2020 | .................... | .................... | ..................... | .................... | .................... | .................... |
| A | 5393 | hCoV-19/Netherlands/NA_26/2020\|EPI_ISL_415483\|2020 | .................... | .................... | ..................... | .................... | .................... | .................... |
| A | 5394 | hCoV-19/Netherlands/NA_27/2020\|EPI_ISL_415484\|2020 | .................... | .................... | ..................... | .................... | .................... | .................... |
| A | 5395 | hCoV-19/Netherlands/NA_28/2020\|EPI_ISL_415485\|2020 | .................... | .................... | ..................... | .................... | .................... | .................... |
| A | 5396 | hCoV-19/Netherlands/NA_29/2020\|EPI_ISL_415486\|2020 | .................... | .................... | ..................... | .................... | .................... | .................... |
| A | 5397 | hCoV-19/Netherlands/NA_30/2020\|EPI_ISL_415487\|2020 | .................... | .................... | ..................... | .................... | .................... | .................... |
| A | 5398 | hCoV-19/Netherlands/NA_31/2020\|EPI_ISL_415488\|2020 | .................... | .................... | ..................... | .................... | .................... | .................... |
| A | 5399 | hCoV-19/Netherlands/NA_32/2020\|EPI_ISL_415489\|2020 | .................... | .................... | ..................... | .................... | .................... | .................... |
| A | 5400 | hCoV-19/Netherlands/NA_34/2020\|EPI_ISL_415491\|2020 | .................... | .................... | ..................... | .................... | .................... | .................... |
| A | 5401 | hCoV-19/Netherlands/NA_35/2020\|EPI_ISL_415492\|2020 | .................... | .................... | ..................... | .................... | .................... | .................... |
| A | 5402 | hCoV-19/Netherlands/NA_4/2020\|EPI_ISL_415493\|2020 | .................... | .................... | ..................... | .................... | .................... | .................... |
| A | 5403 | hCoV-19/Netherlands/NA_5/2020\|EPI_ISL_415494\|2020 | .................... | .................... | ..................... | .................... | .................... | .................... |
| A | 5404 | hCoV-19/Netherlands/NA_6/2020\|EPI_ISL_415495\|2020- | .................... | .................... | ..................... | .................... | .................... | .................... |
| A | 5405 | hCoV-19/Netherlands/NA_7/2020\|EPI_ISL_415496\|2020- | .................... | .................... | ..................... | .................... | .................... | .................... |
| A | 5406 | hCoV-19/Netherlands/NA_8/2020\|EPI_ISL_415497\|2020- | .................... | .................... | ..................... | .................... | .................... | .................... |
| A | 5407 | hCoV-19/Netherlands/NA_9/2020\|EPI_ISL_415498\|2020- | .................... | .................... | ..................... | .................... | .................... | .................... |
| A | 5408 | hCoV-19/Netherlands/NoordBrabant_41/2020\|EPI_ISL_4 | .................... | .................... | ..................... | .................... | .................... | .................... |
| A | 5409 | hCoV-19/Singapore/21/2020\|EPI_ISL_419000\|2020-02-1 | .................... | .................... | ..................... | .................... | .................... | .................... |
| A | 5410 | hCoV-19/Singapore/19/2020\|EPI_ISL_419001\|2020-03-0 | .................... | .................... | ..................... | .................... | .................... | .................... |
| A | 5411 | hCoV-19/Australia/VIC309/2020\|EPI_ISL_420000\|2020- | .................... | .................... | ..................... | .................... | .................... | .................... |
| A | 5412 | hCoV-19/Australia/VIC310/2020\|EPI_ISL_420001\|2020- | .................... | .................... | ..................... | .................... | .................... | .................... |
| A | 5413 | hCoV-19/Australia/VIC311/2020\|EPI_ISL_420002\|2020- | .................... | .................... | ..................... | .................... | .................... | .................... |
| A | 5414 | hCoV-19/Australia/VIC312/2020\|EPI_ISL_420003\|2020- | .................... | .................... | ..................... | .................... | .................... | .................... |
| A | 5415 | hCoV-19/Australia/VIC313/2020\|EPI_ISL_420004\|2020- | .................... | .................... | ..................... | .................... | .................... | .................... |
| A | 5416 | hCoV-19/Australia/VIC314/2020\|EPI_ISL_420005\|2020- | .................... | .................... | ..................... | .................... | .................... | .................... |
| A | 5417 | hCoV-19/Australia/VIC315/2020\|EPI_ISL_420006\|2020- | .................... | .................... | ..................... | .................... | .................... | .................... |

A 5418 hCoV-19/Australia/VIC316/2020|EPI_ISL_420007|2020- ···················· ···················· ···················· ···················· ···················· ····················
A 5419 hCoV-19/Australia/VIC317/2020|EPI_ISL_420008|2020- ···················· ···················· ···················· ···················· ···················· ····················
A 5420 hCoV-19/Australia/VIC318/2020|EPI_ISL_420009|2020- ···················· ···················· ···················· ···················· ···················· ····················
A 5421 hCoV-19/Australia/VIC319/2020|EPI_ISL_420010|2020- ···················· ···················· ···················· ···················· ···················· ····················
A 5422 hCoV-19/Australia/VIC320/2020|EPI_ISL_420011|2020- ···················· ···················· ···················· ···················· ···················· ····················
A 5423 hCoV-19/Australia/VIC321/2020|EPI_ISL_420012|2020- ···················· ···················· ···················· ···················· ···················· ····················
A 5424 hCoV-19/Australia/VIC322/2020|EPI_ISL_420013|2020- ···················· ···················· ···················· ···················· ···················· ····················
A 5425 hCoV-19/Australia/VIC323/2020|EPI_ISL_420014|2020- ···················· ···················· ···················· ···················· ···················· ····················
A 5426 hCoV-19/USA/WI-UW-44/2020|EPI_ISL_421302|2020-03-1 ···················· ···················· ···················· ···················· ···················· ····················
A 5427 hCoV-19/USA/WI-UW-45/2020|EPI_ISL_421303|2020-03-2 ···················· ···················· ···················· ···················· ···················· ····················
A 5428 hCoV-19/USA/WI-UW-46/2020|EPI_ISL_421304|2020-03-2 ···················· ···················· ···················· ···················· ···················· ····················
A 5429 hCoV-19/USA/WI-UW-47/2020|EPI_ISL_421305|2020-03-1 ···················· ···················· ···················· ···················· ···················· ····················
A 5430 hCoV-19/USA/WI-UW-48/2020|EPI_ISL_421306|2020-03-2 ···················· ···················· ···················· ···················· ···················· ····················
A 5431 hCoV-19/USA/WI-UW-49/2020|EPI_ISL_421307|2020-03-2 ···················· ···················· ···················· ···················· ···················· ····················
A 5432 hCoV-19/USA/WI-UW-50/2020|EPI_ISL_421308|2020-03-2 ···················· ···················· ···················· ···················· ···················· ····················
A 5433 hCoV-19/USA/WI-UW-51/2020|EPI_ISL_421309|2020-03-2 ···················· ···················· ···················· ···················· ···················· ····················
A 5434 hCoV-19/USA/WI-UW-52/2020|EPI_ISL_421310|2020-03-1 ···················· ···················· ···················· ···················· ···················· ····················
A 5435 hCoV-19/USA/WI-UW-53/2020|EPI_ISL_421311|2020-03-1 ···················· ···················· ···················· ···················· ···················· ····················
A 5436 hCoV-19/USA/WI-UW-54/2020|EPI_ISL_421312|2020-03-2 ···················· ···················· ···················· ···················· ···················· ····················
A 5437 hCoV-19/USA/WI-UW-55/2020|EPI_ISL_421313|2020-03-2 ···················· ···················· ···················· ···················· ···················· ····················
A 5438 hCoV-19/USA/WI-UW-56/2020|EPI_ISL_421314|2020-03-1 ···················· ···················· ···················· ···················· ···················· ····················
A 5439 hCoV-19/USA/WI-UW-57/2020|EPI_ISL_421315|2020-03-1 ···················· ···················· ···················· ···················· ···················· ····················
A 5440 hCoV-19/USA/WI-UW-58/2020|EPI_ISL_421316|2020-03-2 ···················· ···················· ···················· ···················· ···················· ····················
A 5441 hCoV-19/USA/WI-UW-59/2020|EPI_ISL_421317|2020-03-2 ···················· ···················· ···················· ···················· ···················· ····················
A 5442 hCoV-19/USA/WI-UW-60/2020|EPI_ISL_421318|2020-03-1 ···················· ···················· ···················· ···················· ···················· ····················
A 5443 hCoV-19/USA/WI-UW-61/2020|EPI_ISL_421319|2020-03-2 ···················· ···················· ···················· ···················· ···················· ····················
A 5444 hCoV-19/USA/WI-UW-62/2020|EPI_ISL_421320|2020-03-2 ···················· ···················· ···················· ···················· ···················· ····················
A 5445 hCoV-19/USA/WI-UW-63/2020|EPI_ISL_421321|2020-03-2 ···················· ···················· ···················· ···················· ···················· ····················
A 5446 hCoV-19/USA/WI-UW-64/2020|EPI_ISL_421322|2020-03-2 ···················· ···················· ···················· ···················· ···················· ····················
A 5447 hCoV-19/USA/WI-UW-65/2020|EPI_ISL_421323|2020-03-2 ···················· ···················· ···················· ···················· ···················· ····················
A 5448 hCoV-19/USA/WI-UW-66/2020|EPI_ISL_421324|2020-03-2 ···················· ···················· ···················· ···················· ···················· N···················
A 5449 hCoV-19/USA/WI-UW-67/2020|EPI_ISL_421325|2020-03-2 ···················· ···················· ···················· ···················· ···················· ····················
A 5450 hCoV-19/USA/WI-UW-68/2020|EPI_ISL_421326|2020-03-2 ···················· ···················· ···················· ···················· ···················· ····················
A 5451 hCoV-19/USA/WI-UW-69/2020|EPI_ISL_421327|2020-03-1 ···················· ···················· ···················· ···················· ···················· ····················
A 5452 hCoV-19/USA/WI-UW-70/2020|EPI_ISL_421328|2020-03-1 ···················· ···················· ···················· ···················· ···················· ····················
A 5453 hCoV-19/USA/WI-UW-71/2020|EPI_ISL_421329|2020-03-2 ···················· ···················· ···················· ···················· ···················· ····················
A 5454 hCoV-19/USA/WI-UW-72/2020|EPI_ISL_421330|2020-03-2 ···················· ···················· ···················· ···················· ···················· ····················
A 5455 hCoV-19/USA/WI-UW-73/2020|EPI_ISL_421331|2020-03-2 ···················· ···················· ···················· ···················· ···················· ····················
A 5456 hCoV-19/USA/WI-UW-74/2020|EPI_ISL_421332|2020-03-2 ···················· ···················· ···················· ···················· ···················· ····················
A 5457 hCoV-19/USA/WI-UW-75/2020|EPI_ISL_421333|2020-03-2 ···················· ···················· ···················· ···················· ···················· ····················
A 5458 hCoV-19/USA/WI-UW-76/2020|EPI_ISL_421334|2020-03-2 ···················· ···················· ···················· ···················· ···················· ····················
A 5459 hCoV-19/USA/WI-UW-77/2020|EPI_ISL_421335|2020-03-1 ···················· ···················· ···················· ···················· ···················· ····················
A 5460 hCoV-19/USA/WI-UW-78/2020|EPI_ISL_421336|2020-03-2 ···················· ···················· ···················· ···················· ···················· ····················
A 5461 hCoV-19/USA/WI-UW-79/2020|EPI_ISL_421338|2020-03-2 ···················· ···················· ···················· ···················· ···················· ····················
A 5462 hCoV-19/USA/WI-UW-80/2020|EPI_ISL_421339|2020-03-2 ···················· ···················· ···················· ···················· ···················· ····················
A 5463 hCoV-19/USA/WI-UW-81/2020|EPI_ISL_421340|2020-03-2 ···················· ···················· ···················· ···················· ···················· ····················
A 5464 hCoV-19/USA/WI-UW-82/2020|EPI_ISL_421341|2020-03-2 ···················· ···················· ···················· ···················· ···················· ····················
A 5465 hCoV-19/USA/WI-UW-83/2020|EPI_ISL_421342|2020-03-2 ···················· ···················· ···················· ···················· ···················· ····················
A 5466 hCoV-19/USA/WI-UW-84/2020|EPI_ISL_421343|2020-03-2 ···················· ···················· ···················· ···················· ···················· ····················
A 5467 hCoV-19/Netherlands/NA_302/2020|EPI_ISL_422611|202 ···················· ···················· ···················· ···················· ···················· ····················
A 5468 hCoV-19/Netherlands/NA_303/2020|EPI_ISL_422612|202 ···················· ···········A········ ···················· ···················· ···················· ····················
A 5469 hCoV-19/Netherlands/NA_306/2020|EPI_ISL_422615|202 ···················· ···················· ···················· ···················· ···················· ····················
A 5470 hCoV-19/Netherlands/NA_307/2020|EPI_ISL_422616|202 ···················· ···················· ···················· ···················· ···················· ····················
A 5471 hCoV-19/Netherlands/NA_311/2020|EPI_ISL_422620|202 ···················· ···················· ···················· ···················· ···················· ····················
A 5472 hCoV-19/Netherlands/NA_312/2020|EPI_ISL_422621|202 ···················· ···················· ···················· ···················· ···················· ····················
A 5473 hCoV-19/Netherlands/NA_314/2020|EPI_ISL_422623|202 ···················· ···················· ···················· ···················· ···················· ····················
A 5474 hCoV-19/Netherlands/ZuidHolland_97/2020|EPI_ISL_42 ···················· ···················· ···················· ···················· ···················· ····················
A 5475 hCoV-19/Netherlands/ZuidHolland_98/2020|EPI_ISL_42 ···················· ···················· ···················· ···················· ···················· ····················
A 5476 hCoV-19/Netherlands/ZuidHolland_99/2020|EPI_ISL_42 ···················· ···················· ···················· ···················· ···················· ····················
A 5477 hCoV-19/Czech Republic/2741/2020|EPI_ISL_422636|20 ···················· ···················· ···················· ···················· ···················· ····················
A 5478 hCoV-19/Netherlands/Gelderland_10/2020|EPI_ISL_422 ···················· ···················· ···················· ···················· ···················· ····················
A 5479 hCoV-19/Netherlands/Gelderland_11/2020|EPI_ISL_422 ···················· ···················· ···················· ···················· ···················· ····················
A 5480 hCoV-19/Netherlands/Gelderland_12/2020|EPI_ISL_422 ···················· ···················· ···················· ···················· ···················· ····················
A 5481 hCoV-19/Netherlands/Gelderland_5/2020|EPI_ISL_4226 ···················· ···················· ···················· ···················· ···················· ····················
A 5482 hCoV-19/Netherlands/Gelderland_6/2020|EPI_ISL_4226 ···················· ···················· ···················· ···················· ···················· ····················
A 5483 hCoV-19/Netherlands/Gelderland_7/2020|EPI_ISL_4226 ···················· ···················· ···················· ···················· ···················· ····················
A 5484 hCoV-19/Netherlands/Gelderland_8/2020|EPI_ISL_4226 ···················· ···················· ···················· ···················· ···················· ····················
A 5485 hCoV-19/Netherlands/Gelderland_9/2020|EPI_ISL_4226 ···················· ···················· ···················· ···················· ···················· ····················
A 5486 hCoV-19/Netherlands/Limburg_10/2020|EPI_ISL_422646 ···················· ···················· ···················· ···················· ···················· ····················
A 5487 hCoV-19/Netherlands/Limburg_8/2020|EPI_ISL_422647| ···················· ···················· ···················· ···················· ···················· ····················
A 5488 hCoV-19/Netherlands/Limburg_9/2020|EPI_ISL_422648| ···················· ···················· ···················· ···················· ···················· ····················
A 5489 hCoV-19/Netherlands/NA_100/2020|EPI_ISL_422649|202 ···················· ···················· ···················· ···················· ···················· ····················
A 5490 hCoV-19/Netherlands/NA_101/2020|EPI_ISL_422650|202 ···················· ···················· ···················· ···················· ···················· ····················
A 5491 hCoV-19/Netherlands/NA_102/2020|EPI_ISL_422651|202 ···················· ···················· ···················· ···················· ···················· ····················
A 5492 hCoV-19/Netherlands/NA_103/2020|EPI_ISL_422652|202 ···················· ···················· ···················· ···················· ···················· ····················
A 5493 hCoV-19/Netherlands/NA_104/2020|EPI_ISL_422653|202 ···················· ···················· ···················· ···················· ···················· ····················
A 5494 hCoV-19/Netherlands/NA_106/2020|EPI_ISL_422655|202 ···················· ···················· ···················· ···················· ···················· ····················
A 5495 hCoV-19/Netherlands/NA_107/2020|EPI_ISL_422656|202 ···················· ···················· ···················· ···················· ···················· ····················

A 5496 hCoV-19/Netherlands/NA_109/2020|EPI_ISL_422657|202 ···················· ···················· ···················· ···················· ···················· ····················
A 5497 hCoV-19/Netherlands/NA_111/2020|EPI_ISL_422659|202 ···················· ···················· ···················· ···················· ···················· ····················
A 5498 hCoV-19/Netherlands/NA_112/2020|EPI_ISL_422660|202 ···················· ···················· ···················· ···················· ···················· ····················
A 5499 hCoV-19/Netherlands/NA_113/2020|EPI_ISL_422661|202 ···················· ···················· ···················· ···················· ···················· ····················
A 5500 hCoV-19/Netherlands/NA_114/2020|EPI_ISL_422662|202 ···················· ···················· ···················· ···················· ···················· ····················
A 5501 hCoV-19/Netherlands/NA_117/2020|EPI_ISL_422664|202 ···················· ···················· ···················· ···················· ···T················ ····················
A 5502 hCoV-19/Netherlands/NA_118/2020|EPI_ISL_422665|202 ···················· ···················· ···················· ···················· ···T················ ····················
A 5503 hCoV-19/Netherlands/NA_119/2020|EPI_ISL_422666|202 ···················· ···················· ···················· ···················· ···················· ····················
A 5504 hCoV-19/Netherlands/NA_120/2020|EPI_ISL_422667|202 ···················· ···················· ···················· ···················· ···················· ····················
A 5505 hCoV-19/Netherlands/NA_121/2020|EPI_ISL_422668|202 ···················· ···················· ···················· ···················· ···················· ····················
A 5506 hCoV-19/Netherlands/NA_122/2020|EPI_ISL_422669|202 ···················· ···················· ···················· ···················· ···················· ····················
A 5507 hCoV-19/Netherlands/NA_126/2020|EPI_ISL_422670|202 ···················· ···················· ···················· ···················· ···················· ····················
A 5508 hCoV-19/Netherlands/NA_127/2020|EPI_ISL_422671|202 ···················· ···················· ···················· ···················· ···················· ····················
A 5509 hCoV-19/Netherlands/NA_128/2020|EPI_ISL_422672|202 ···················· ···················· ···················· ···················· ···················· ····················
A 5510 hCoV-19/Netherlands/NA_129/2020|EPI_ISL_422673|202 ···················· ···················· ···················· ···················· ···················· ····················
A 5511 hCoV-19/Netherlands/NA_130/2020|EPI_ISL_422674|202 ···················· ···················· ···················· ···················· ···················· ····················
A 5512 hCoV-19/Netherlands/NA_131/2020|EPI_ISL_422675|202 ···················· ···················· ···················· ···················· ···················· ····················
A 5513 hCoV-19/Netherlands/NA_132/2020|EPI_ISL_422676|202 ···················· ···················· ···················· ···················· ···················· ····················
A 5514 hCoV-19/England/20126097804/2020|EPI_ISL_423906|20 ···················· ···················· ···················· ···················· ···················· ····················
A 5515 hCoV-19/England/20126100404/2020|EPI_ISL_423907|20 ···················· ···················· ···················· ···················· ···················· ····················
A 5517 hCoV-19/England/20126103304/2020|EPI_ISL_423911|20 ···················· ···················· ···················· ···················· ···················· ····················
A 5518 hCoV-19/England/20128000604/2020|EPI_ISL_423918|20 ···················· ···················· ···················· ···················· ···················· ····················
A 5519 hCoV-19/England/20128015104/2020|EPI_ISL_423942|20 ···················· ···················· ···················· ···················· ···················· ····················
A 5520 hCoV-19/England/20128015604/2020|EPI_ISL_423943|20 ···················· ···················· ···················· ···················· ···················· ····················
A 5521 hCoV-19/England/20128018804/2020|EPI_ISL_423948|20 ···················· ···················· ···················· ···················· ···················· ····················
A 5523 hCoV-19/England/20128036004/2020|EPI_ISL_423952|20 ···················· ···················· ···················· ···················· ···················· ····················
A 5524 hCoV-19/England/20128085204/2020|EPI_ISL_423964|20 ···················· ···················· ···················· ···················· ···················· ····················
A 5525 hCoV-19/England/20128090404/2020|EPI_ISL_423965|20 ···················· ···················· ···················· ···················· ···················· ····················
A 5527 hCoV-19/England/20128094904/2020|EPI_ISL_423969|20 ···················· ···················· ···················· ···················· ···················· ····················
A 5529 hCoV-19/England/20129000404/2020|EPI_ISL_423973|20 ···················· ···················· ···················· ···················· ···················· ····················
A 5530 hCoV-19/England/20129002004/2020|EPI_ISL_423976|20 ···················· ···················· ···················· ···················· ···················· ····················
A 5531 hCoV-19/England/20129009304/2020|EPI_ISL_423979|20 ···················· ···················· ···················· ···················· ···················· ····················
A 5532 hCoV-19/England/20129012304/2020|EPI_ISL_423990|20 ···················· ···················· ···················· ···················· ···················· ····················
A 5533 hCoV-19/England/20129015404/2020|EPI_ISL_423998|20 ···················· ···················· ···················· ···················· ···················· ····················
A 5534 hCoV-19/England/20129015704/2020|EPI_ISL_423999|20 ···················· ···················· ···················· ···················· ···················· ····················
A 5535 hCoV-19/USA/WA-UW-2230/2020|EPI_ISL_426120|2020-03 ···················· ···················· ···················· ···················· ···················· ····················
A 5536 hCoV-19/USA/WA-UW-2232/2020|EPI_ISL_426121|2020-03 ···················· ···················· ···················· ···················· ···················· ····················
A 5537 hCoV-19/USA/WA-UW-2234/2020|EPI_ISL_426122|2020-03 ···················· ···················· ···················· ···················· ···················· ····················
A 5538 hCoV-19/USA/WA-UW-2235/2020|EPI_ISL_426123|2020-03 ···················· ···················· ···················· ···················· ···················· ····················
A 5539 hCoV-19/USA/WA-UW-2236/2020|EPI_ISL_426124|2020-03 ···················· ···················· ···················· ···················· ···················· ····················
A 5540 hCoV-19/USA/WA-UW-2238/2020|EPI_ISL_426125|2020-03 ···················· ···················· ···················· ···················· ···················· ····················
A 5541 hCoV-19/USA/WA-UW-2240/2020|EPI_ISL_426126|2020-03 ···················· ···················· ···················· ···················· ···················· ····················
A 5542 hCoV-19/USA/WA-UW-2244/2020|EPI_ISL_426127|2020-03 ···················· ···················· ···················· ···················· ···················· ····················
A 5543 hCoV-19/USA/WA-UW-2245/2020|EPI_ISL_426128|2020-03 ···················· ···················· ···················· ···················· ···················· ····················
A 5544 hCoV-19/USA/WA-UW-2247/2020|EPI_ISL_426129|2020-03 ···················· ···················· ···················· ···················· ···················· ····················
A 5545 hCoV-19/USA/WA-UW-2249/2020|EPI_ISL_426130|2020-03 ···················· ···················· ···················· ···················· ···················· ····················
A 5546 hCoV-19/USA/WA-UW-2250/2020|EPI_ISL_426131|2020-03 ···················· ···················· ···················· ···················· ···················· ····················
A 5547 hCoV-19/USA/WA-UW-2253/2020|EPI_ISL_426132|2020-03 ···················· ···················· ···················· ···················· ···················· ····················
A 5548 hCoV-19/USA/WA-UW-2258/2020|EPI_ISL_426133|2020-03 ···················· ···················· ···················· ···················· ···················· ····················
A 5549 hCoV-19/USA/WA-UW-4404/2020|EPI_ISL_426135|2020-03 ···················· ···················· ···················· ···················· ···················· ····················
A 5550 hCoV-19/USA/WA-UW-4406/2020|EPI_ISL_426136|2020-03 ···················· ···················· ···················· ···················· ···················· ····················
A 5551 hCoV-19/USA/WA-UW-4407/2020|EPI_ISL_426137|2020-03 ···················· ···················· ···················· ···················· ···················· ····················
A 5552 hCoV-19/USA/WI-GMF-00986/2020|EPI_ISL_426159|2020- ···················· ···················· ···················· ···················· ···················· ····················
A 5553 hCoV-19/USA/WI-GMF-00921/2020|EPI_ISL_426160|2020- ···················· ···················· ···················· ···················· ···················· ····················
A 5554 hCoV-19/USA/WI-GMF-00928/2020|EPI_ISL_426161|2020- ···················· ···················· ···················· ···················· ···················· ····················
A 5555 hCoV-19/South Korea/KCDC2003/2020|EPI_ISL_426163|2 ···················· ···················· ···················· ···················· ···················· ····················
A 5556 hCoV-19/South Korea/KCDC2004/2020|EPI_ISL_426164|2 ···················· ···················· ···················· ···················· ···················· ····················
A 5557 hCoV-19/South Korea/KCDC2005/2020|EPI_ISL_426166|2 ···················· ···················· ···················· ···················· ···················· ····················
A 5558 hCoV-19/South Korea/KCDC2003/2020|EPI_ISL_426167|2 ···················· ···················· ···················· ···················· ···················· ····················
A 5559 hCoV-19/South Korea/KCDC2006/2020|EPI_ISL_426168|2 ···················· ···················· ···················· ···················· ···················· ····················
A 5560 hCoV-19/South Korea/KCDC2007/2020|EPI_ISL_426169|2 ···················· ···················· ···················· ···················· ···················· ····················
A 5561 hCoV-19/South Korea/KCDC2008/2020|EPI_ISL_426171|2 ···················· ···················· ···················· ···················· ···················· ····················
A 5562 hCoV-19/South Korea/KCDC2009/2020|EPI_ISL_426173|2 ···················· ···················· ···················· ···················· ···················· ····················
A 5563 hCoV-19/South Korea/KCDC2010/2020|EPI_ISL_426180|2 ···················· ···················· ···················· ···················· ···················· ····················
A 5564 hCoV-19/South Korea/KCDC2011/2020|EPI_ISL_426181|2 ···················· ···················· ···················· ···················· ···················· ····················
A 5565 hCoV-19/South Korea/KCDC2012/2020|EPI_ISL_426182|2 ···················· ···················· ···················· ···················· ···················· ····················
A 5566 hCoV-19/South Korea/KCDC2013/2020|EPI_ISL_426183|2 ···················· ···················· ···················· ···················· ···················· ····················
A 5567 hCoV-19/South Korea/KCDC2014/2020|EPI_ISL_426187|2 ···················· ···················· ···················· ···················· ···················· ····················
A 5568 hCoV-19/USA/IL-UW-143/2020|EPI_ISL_427450|2020-04- ···················· ···················· ···················· ···················· ···················· ····················
A 5569 hCoV-19/USA/WI-UW-144/2020|EPI_ISL_427451|2020-04- ···················· ···················· ···················· ···················· ···················· ····················
A 5570 hCoV-19/USA/WI-UW-145/2020|EPI_ISL_427452|2020-04- ···················· ···················· ···················· ···················· ···················· ····················
A 5571 hCoV-19/USA/WI-UW-146/2020|EPI_ISL_427453|2020-04- ···················· ···················· ···················· ···················· ···················· ····················
A 5572 hCoV-19/USA/WI-UW-147/2020|EPI_ISL_427454|2020-04- ···················· ···················· ···················· ···················· ···················· ····················
A 5573 hCoV-19/USA/WI-UW-148/2020|EPI_ISL_427455|2020-04- ···················· ···················· ···················· ···················· ···················· ····················
A 5574 hCoV-19/USA/IL-UW-149/2020|EPI_ISL_427456|2020-04- ···················· ···················· ···················· ···················· ···················· ····················
A 5575 hCoV-19/USA/WI-UW-150/2020|EPI_ISL_427457|2020-04- ···················· ···················· ···················· ···················· ···················· ····················
A 5576 hCoV-19/USA/WI-UW-151/2020|EPI_ISL_427458|2020-04- ···················· ···················· ···················· ···················· ···················· ····················
A 5577 hCoV-19/USA/WI-UW-152/2020|EPI_ISL_427459|2020-04- ···················· ···················· ···················· ···················· ···················· ····················

| | | | | | | | |
|---|---|---|---|---|---|---|---|
| A | 5578 hCoV-19/USA/WI-UW-153/2020\|EPI_ISL_427460\|2020-04- | ···················· | ···················· | ···················· | ···················· | ···················· | ···················· |
| A | 5579 hCoV-19/USA/WI-UW-154/2020\|EPI_ISL_427461\|2020-04- | ···················· | ···················· | ···················· | ···················· | ···················· | ···················· |
| A | 5580 hCoV-19/USA/WI-UW-155/2020\|EPI_ISL_427462\|2020-04- | ···················· | ···················· | ···················· | ···················· | ···················· | ···················· |
| A | 5581 hCoV-19/USA/NY-NYUMC172/2020\|EPI_ISL_427469\|2020-0 | ···················· | ···················· | ···················· | ···················· | ···················· | ···················· |
| A | 5582 hCoV-19/USA/NY-NYUMC173/2020\|EPI_ISL_427470\|2020-0 | ···················· | ···················· | ···················· | ···················· | ···················· | ···················· |
| A | 5583 hCoV-19/USA/NY-NYUMC174/2020\|EPI_ISL_427471\|2020-0 | ···················· | ···················· | ···················· | ···················· | ···················· | ···················· |
| A | 5584 hCoV-19/USA/NY-NYUMC175/2020\|EPI_ISL_427472\|2020-0 | ···················· | ···················· | ···················· | ···················· | ···················· | ···················· |
| A | 5585 hCoV-19/USA/NY-NYUMC177/2020\|EPI_ISL_427473\|2020-0 | ···················· | ···················· | ···················· | ···················· | ···················· | ···················· |
| A | 5586 hCoV-19/USA/NY-NYUMC178/2020\|EPI_ISL_427474\|2020-0 | ···················· | ···················· | ···················· | ···················· | ···················· | ···················· |
| A | 5587 hCoV-19/USA/NY-NYUMC179/2020\|EPI_ISL_427475\|2020-0 | ···················· | ···················· | ···················· | ···················· | ···················· | ···················· |
| A | 5588 hCoV-19/USA/NY-NYUMC180/2020\|EPI_ISL_427476\|2020-0 | ···················· | ···················· | ···················· | ···················· | ···················· | ···················· |
| A | 5589 hCoV-19/USA/NY-NYUMC181/2020\|EPI_ISL_427477\|2020-0 | ···················· | ···················· | ···················· | ···················· | ···················· | ···················· |
| A | 5590 hCoV-19/USA/NY-NYUMC182/2020\|EPI_ISL_427478\|2020-0 | ···················· | ···················· | ···················· | ···················· | ···················· | ···················· |
| A | 5591 hCoV-19/USA/NY-NYUMC183/2020\|EPI_ISL_427479\|2020-0 | ···················· | ···················· | ···················· | ···················· | ···················· | ···················· |
| A | 5592 hCoV-19/USA/NY-NYUMC184/2020\|EPI_ISL_427480\|2020-0 | ···················· | ···················· | ···················· | ···················· | ···················· | ···················· |
| A | 5593 hCoV-19/USA/NY-NYUMC185/2020\|EPI_ISL_427481\|2020-0 | ···················· | ···················· | ···················· | ···················· | ···················· | ···················· |
| A | 5594 hCoV-19/USA/NY-NYUMC186/2020\|EPI_ISL_427482\|2020-0 | ···················· | ···················· | ···················· | ···················· | ···················· | ······G············· |
| A | 5595 hCoV-19/USA/NY-NYUMC188/2020\|EPI_ISL_427484\|2020-0 | ···················· | ···················· | ···················· | ···················· | ···················· | ···················· |
| A | 5596 hCoV-19/USA/NY-NYUMC189/2020\|EPI_ISL_427485\|2020-0 | ···················· | ···················· | ···················· | ···················· | ···················· | ···················· |
| A | 5597 hCoV-19/USA/NY-NYUMC192/2020\|EPI_ISL_427487\|2020-0 | ···················· | ···················· | ···················· | ···················· | ···················· | ···················· |
| A | 5598 hCoV-19/USA/NY-NYUMC193/2020\|EPI_ISL_427488\|2020-0 | ···················· | ···················· | ···················· | ···················· | ···················· | ···················· |
| A | 5599 hCoV-19/USA/NY-NYUMC194/2020\|EPI_ISL_427489\|2020-0 | ···················· | ···················· | ···················· | ···················· | ···················· | ···················· |
| A | 5600 hCoV-19/USA/NY-NYUMC195/2020\|EPI_ISL_427490\|2020-0 | ···················· | ···················· | ···················· | ···················· | ···················· | ···················· |
| A | 5601 hCoV-19/USA/NY-NYUMC196/2020\|EPI_ISL_427491\|2020-0 | ···················· | ···················· | ···················· | ···················· | ···················· | ···················· |
| A | 5602 hCoV-19/USA/NY-NYUMC197/2020\|EPI_ISL_427492\|2020-0 | ···················· | ···················· | ···················· | ···················· | ···················· | ···················· |
| A | 5603 hCoV-19/USA/NY-NYUMC198/2020\|EPI_ISL_427493\|2020-0 | ···················· | ···················· | ···················· | ···················· | ···················· | ···················· |
| A | 5604 hCoV-19/USA/NY-NYUMC199/2020\|EPI_ISL_427494\|2020-0 | ···················· | ···················· | ···················· | ···················· | ···················· | ···················· |
| A | 5605 hCoV-19/USA/NY-NYUMC200/2020\|EPI_ISL_427495\|2020-0 | ···················· | ···················· | ···················· | ···················· | ···················· | ···················· |
| A | 5606 hCoV-19/USA/NY-NYUMC201/2020\|EPI_ISL_427496\|2020-0 | ···················· | ···················· | ···················· | ···················· | ···················· | ···················· |
| A | 5607 hCoV-19/USA/NY-NYUMC203/2020\|EPI_ISL_427498\|2020-0 | ···················· | ···················· | ···················· | ···················· | ···················· | ···················· |
| A | 5608 hCoV-19/USA/NY-NYUMC266/2020\|EPI_ISL_428780\|2020-0 | ···················· | ···················· | ···················· | ···················· | ···················· | ···················· |
| A | 5609 hCoV-19/USA/NY-NYUMC267/2020\|EPI_ISL_428781\|2020-0 | ···················· | ···················· | ···················· | ···················· | ···················· | ···················· |
| A | 5610 hCoV-19/USA/NY-NYUMC268/2020\|EPI_ISL_428782\|2020-0 | ···················· | ···················· | ···················· | ···················· | ···················· | ···················· |
| A | 5611 hCoV-19/USA/NY-NYUMC269/2020\|EPI_ISL_428783\|2020-0 | ···················· | ···················· | ···················· | ···················· | ···················· | ···················· |
| A | 5612 hCoV-19/USA/NY-NYUMC270/2020\|EPI_ISL_428784\|2020-0 | ···················· | ···················· | ···················· | ···················· | ···················· | ···················· |
| A | 5613 hCoV-19/USA/NY-NYUMC271/2020\|EPI_ISL_428785\|2020-0 | ···················· | ···················· | ···················· | ···················· | ···················· | ···················· |
| A | 5614 hCoV-19/USA/NY-NYUMC272/2020\|EPI_ISL_428786\|2020-0 | ···················· | ···················· | ···················· | ···················· | ···················· | ···················· |
| A | 5615 hCoV-19/USA/NY-NYUMC273/2020\|EPI_ISL_428787\|2020-0 | ···················· | ···················· | ···················· | ···················· | ···················· | ···················· |
| A | 5616 hCoV-19/USA/NY-NYUMC274/2020\|EPI_ISL_428788\|2020-0 | ···················· | ···················· | ···················· | ···················· | ···················· | ···················· |
| A | 5617 hCoV-19/USA/NY-NYUMC275/2020\|EPI_ISL_428789\|2020-0 | ···················· | ···················· | ···················· | ···················· | ···················· | ···················· |
| A | 5618 hCoV-19/USA/NY-NYUMC276/2020\|EPI_ISL_428790\|2020-0 | ···················· | ···················· | ···················· | ···················· | ···················· | ···················· |
| A | 5619 hCoV-19/USA/NY-NYUMC277/2020\|EPI_ISL_428791\|2020-0 | ···················· | ···················· | ···················· | ···················· | ···················· | ···················· |
| A | 5620 hCoV-19/USA/NY-NYUMC278/2020\|EPI_ISL_428792\|2020-0 | ···················· | ···················· | ···················· | ···················· | ···················· | ···················· |
| A | 5621 hCoV-19/USA/NY-NYUMC279/2020\|EPI_ISL_428793\|2020-0 | ···················· | ···················· | ···················· | ···················· | ···················· | ···················· |
| A | 5622 hCoV-19/USA/NY-NYUMC281/2020\|EPI_ISL_428795\|2020-0 | ···················· | ···················· | ···················· | ···················· | ···················· | ···················· |
| A | 5623 hCoV-19/USA/NY-NYUMC283/2020\|EPI_ISL_428797\|2020-0 | ···················· | ···················· | ···················· | ···················· | ···················· | ···················· |
| A | 5624 hCoV-19/USA/NY-NYUMC284/2020\|EPI_ISL_428798\|2020-0 | ···················· | ···················· | ···················· | ···················· | ···················· | ···················· |
| A | 5625 hCoV-19/USA/NY-NYUMC285/2020\|EPI_ISL_428799\|2020-0 | ···················· | ···················· | ···················· | ···················· | ···················· | ···················· |
| A | 5626 hCoV-19/USA/WA-UW-328/2020\|EPI_ISL_524437\|2020-03- | ···················· | ···················· | ···················· | ···················· | ···················· | ···················· |
| A | 5627 hCoV-19/USA/WA-UW-334/2020\|EPI_ISL_524438\|2020-03- | ···················· | ···················· | ···················· | ···················· | ···················· | ···················· |
| A | 5628 hCoV-19/USA/WA-UW-350/2020\|EPI_ISL_524439\|2020-03- | ···················· | ···················· | ···················· | ···················· | ···················· | ···················· |
| A | 5629 hCoV-19/USA/WA-UW-351/2020\|EPI_ISL_524440\|2020-03- | ···················· | ···················· | ···················· | ···················· | ···················· | ···················· |
| A | 5630 hCoV-19/USA/WA-UW-352/2020\|EPI_ISL_524441\|2020-03- | ···················· | ···················· | ···················· | ···················· | ···················· | ···················· |
| A | 5631 hCoV-19/USA/WA-UW-354/2020\|EPI_ISL_524442\|2020-03- | ···················· | ···················· | ···················· | ···················· | ···················· | ···················· |
| A | 5632 hCoV-19/USA/WA-UW-380/2020\|EPI_ISL_524443\|2020-03- | ···················· | ···················· | ···················· | ···················· | ···················· | ···················· |
| A | 5633 hCoV-19/Wuhan/IVDC-HB-01/2019\|EPI_ISL_402119\|2019- | ···················· | ···················· | ···················· | ···················· | ···················· | ···················· |
| A | 5634 hCoV-19/Wuhan/IVDC-HB-04/2020\|EPI_ISL_402120\|2020- | ···················· | ···················· | ···················· | ···················· | ···················· | ···················· |
| A | 5635 hCoV-19/Wuhan/IVDC-HB-05/2019\|EPI_ISL_402121\|2019- | ···················· | ···················· | ···················· | ···················· | ···················· | ···················· |
| A | 5636 hCoV-19/Wuhan/IPBCAMS-WH-01/2019\|EPI_ISL_402123\|20 | ···················· | ···················· | ···················· | ···················· | ···················· | ···················· |
| A | 5637 hCoV-19/Wuhan/WIV04/2019\|EPI_ISL_402124\|2019-12-30 | ···················· | ···················· | ···················· | ···················· | ···················· | ···················· |
| A | 5638 hCoV-19/Wuhan/Hu-1/2019\|EPI_ISL_402125\|2019-12-31 | ···················· | ···················· | ···················· | ···················· | ···················· | ···················· |
| A | 5639 hCoV-19/Wuhan/WIV02/2019\|EPI_ISL_402127\|2019-12-30 | ···················· | ···················· | ···················· | ···················· | ···················· | ···················· |
| A | 5640 hCoV-19/Wuhan/WIV05/2019\|EPI_ISL_402128\|2019-12-30 | ···················· | ···················· | ···················· | ···················· | ···················· | ···················· |
| A | 5641 hCoV-19/Wuhan/WIV06/2019\|EPI_ISL_402129\|2019-12-30 | ···················· | ···················· | ···················· | ···················· | ···················· | ···················· |
| A | 5642 hCoV-19/Wuhan/WIV07/2019\|EPI_ISL_402130\|2019-12-30 | ···················· | ···················· | ···················· | ···················· | ···················· | ···················· |
| A | 5643 hCoV-19/Wuhan/HBCDC-HB-01/2019\|EPI_ISL_402132\|2019 | ···················· | ···················· | ···················· | ···················· | ···················· | ···················· |
| A | 5644 hCoV-19/South Korea/SNU01/2020\|EPI_ISL_411929\|2020 | ···················· | ···················· | ···················· | ···········T········ | ···················· | ···················· |
| A | 5645 hCoV-19/Switzerland/GE-HUG-1422/2020\|EPI_ISL_41545 | ···················· | ···················· | ···················· | ···················· | ···················· | ···················· |
| A | 5646 hCoV-19/Switzerland/GE-HUG-0199/2020\|EPI_ISL_41545 | ···················· | ···················· | ···················· | ···················· | ···················· | ···················· |
| A | 5647 hCoV-19/Switzerland/BE-HUG-6651/2020\|EPI_ISL_41545 | ···················· | ···················· | ···················· | ···················· | ···················· | ···················· |
| A | 5648 hCoV-19/Switzerland/AG-HUG-7120/2020\|EPI_ISL_41545 | ···················· | ···················· | ···················· | ···················· | ···················· | ···················· |
| A | 5649 hCoV-19/Switzerland/GE-HUG-8102/2020\|EPI_ISL_41545 | ···················· | ···················· | ···················· | ···················· | ···················· | ···················· |
| A | 5650 hCoV-19/Switzerland/GE-HUG-VD0503/2020\|EPI_ISL_415 | ···················· | ···················· | ···················· | ···················· | ···················· | ···················· |
| A | 5651 hCoV-19/Netherlands/Flevoland_1/2020\|EPI_ISL_41546 | ···················· | ···················· | ···················· | ···················· | ···················· | ···················· |
| A | 5652 hCoV-19/Netherlands/Gelderland_1-2/2020\|EPI_ISL_41 | ···················· | ···················· | ···················· | ···················· | ···················· | ···················· |
| A | 5653 hCoV-19/Netherlands/Gelderland_2/2020\|EPI_ISL_4154 | ···················· | ···················· | ···················· | ···················· | ···················· | ···················· |
| A | 5654 hCoV-19/Netherlands/Gelderland_3/2020\|EPI_ISL_4154 | ···················· | ···················· | ···················· | ···················· | ···················· | ···················· |
| A | 5655 hCoV-19/Netherlands/Limburg_7/2020\|EPI_ISL_415464\| | ···················· | ···················· | ···················· | ···················· | ···················· | ···················· |

| | | | | | | | |
|---|---|---|---|---|---|---|---|
| A | 5656 hCoV-19/Netherlands/NA_1/2020\|EPI_ISL_415465\|2020- | ···················· | ···················· | ···················· | ···················· | ···················· | ···················· |
| A | 5657 hCoV-19/Netherlands/NA_10/2020\|EPI_ISL_415466\|2020 | ···················· | ···················· | ···················· | ···················· | ···················· | ···················· |
| A | 5658 hCoV-19/Netherlands/NA_11/2020\|EPI_ISL_415467\|2020 | ···················· | ···················· | ···················· | ···················· | ···················· | ···················· |
| A | 5659 hCoV-19/Netherlands/NA_12/2020\|EPI_ISL_415468\|2020 | ···················· | ···················· | ···················· | ···················· | ···················· | ···················· |
| A | 5660 hCoV-19/Netherlands/NA_13/2020\|EPI_ISL_415469\|2020 | ···················· | ···················· | ···················· | ···················· | ···················· | ···················· |
| A | 5661 hCoV-19/USA/WA-UW177/2020\|EPI_ISL_416715\|2020-03-1 | ···················· | ···················· | ···················· | ···················· | ···················· | ···················· |
| A | 5662 hCoV-19/USA/WA-UW178/2020\|EPI_ISL_416716\|2020-03-1 | ···················· | ···················· | ···················· | ···················· | ···················· | ···················· |
| A | 5663 hCoV-19/USA/WA-UW179/2020\|EPI_ISL_416717\|2020-03-1 | ···················· | ···················· | ···················· | ···················· | ···················· | ···················· |
| A | 5664 hCoV-19/USA/WA-UW180/2020\|EPI_ISL_416718\|2020-03-1 | ···················· | ···················· | ···················· | ···················· | ···················· | ···················· |
| A | 5665 hCoV-19/USA/WA-UW181/2020\|EPI_ISL_416719\|2020-03-1 | ···················· | ···················· | ···················· | ···················· | ···················· | ···················· |
| A | 5666 hCoV-19/USA/WA-UW183/2020\|EPI_ISL_416721\|2020-03-1 | ···················· | ···················· | ···················· | ···················· | ···················· | ···················· |
| A | 5667 hCoV-19/USA/WA-UW184/2020\|EPI_ISL_416722\|2020-03-1 | ···················· | ···················· | ···················· | ···················· | ···················· | ···················· |
| A | 5668 hCoV-19/USA/WA-UW185/2020\|EPI_ISL_416723\|2020-03-1 | ···················· | ···················· | ···················· | ···················· | ···················· | ···················· |
| A | 5669 hCoV-19/USA/WA-UW186/2020\|EPI_ISL_416724\|2020-03-1 | ···················· | ···················· | ···················· | ···················· | ···················· | ···················· |
| A | 5670 hCoV-19/USA/WA-UW187/2020\|EPI_ISL_416725\|2020-03-1 | ···················· | ···················· | ···················· | ···················· | ···················· | ···················· |
| A | 5671 hCoV-19/USA/WA-UW188/2020\|EPI_ISL_416726\|2020-03-1 | ···················· | ···················· | ···················· | ···················· | ···················· | ···················· |
| A | 5672 hCoV-19/USA/WA-UW189/2020\|EPI_ISL_416727\|2020-03-1 | ···················· | ···················· | ···················· | ···················· | ···················· | ···················· |
| A | 5673 hCoV-19/USA/WA-UW190/2020\|EPI_ISL_416728\|2020-03-1 | ···················· | ···················· | ···················· | ···················· | ···················· | ···················· |
| A | 5674 hCoV-19/USA/WA-UW191/2020\|EPI_ISL_416729\|2020-03-1 | ···················· | ···················· | ···················· | ···················· | ···················· | ···················· |
| A | 5675 hCoV-19/England/SHEF-BFCB1/2020\|EPI_ISL_416730\|202 | ···················· | ···················· | ···················· | ···················· | ···················· | ···················· |
| A | 5676 hCoV-19/England/SHEF-BFCC0/2020\|EPI_ISL_416731\|202 | ···················· | ···················· | ···················· | ···················· | ···················· | ···················· |
| A | 5677 hCoV-19/England/SHEF-BFCDF/2020\|EPI_ISL_416732\|202 | ···················· | ···················· | ···················· | ···················· | ···················· | ···················· |
| A | 5678 hCoV-19/England/SHEF-BFCEE/2020\|EPI_ISL_416733\|202 | ···················· | ···················· | ···················· | ···················· | ···················· | ···················· |
| A | 5679 hCoV-19/England/SHEF-BFCFD/2020\|EPI_ISL_416734\|202 | ···················· | ···················· | ···················· | ···················· | ···················· | ···················· |
| A | 5681 hCoV-19/England/SHEF-BFD18/2020\|EPI_ISL_416736\|202 | ···················· | ···················· | ···················· | ···················· | ···················· | ···················· |
| A | 5683 hCoV-19/England/SHEF-BFD36/2020\|EPI_ISL_416738\|202 | ···················· | ···················· | ···················· | ···················· | ···················· | ···················· |
| A | 5684 hCoV-19/England/SHEF-BFD45/2020\|EPI_ISL_416739\|202 | ···················· | ···················· | ···················· | ···················· | ···················· | ···················· |
| A | 5685 hCoV-19/England/SHEF-BFD54/2020\|EPI_ISL_416740\|202 | ···················· | ···················· | ···················· | ···················· | ···················· | ···················· |
| A | 5686 hCoV-19/Lithuania/ChVir1632/2020\|EPI_ISL_416741\|20 | ···················· | ···················· | ···················· | ···················· | ···················· | ···················· |
| A | 5687 hCoV-19/Czech Republic/ChVir1630/2020\|EPI_ISL_4167 | ···················· | ···················· | ···················· | ···················· | ···················· | ···················· |
| A | 5688 hCoV-19/Czech Republic/ChVir1912/2020\|EPI_ISL_4167 | ···················· | ···················· | ···················· | ···················· | ···················· | ···················· |
| A | 5689 hCoV-19/Hungary/mbl49/2020\|EPI_ISL_416744\|2020-03- | ···················· | ···················· | ···················· | ···················· | ···················· | ···················· |
| A | 5690 hCoV-19/France/ARA-1733/2020\|EPI_ISL_416745\|2020-0 | ···················· | ···················· | ···················· | ···················· | ···················· | ···················· |
| A | 5691 hCoV-19/France/ARA-425/2020\|EPI_ISL_416746\|2020-03 | ···················· | ···················· | ···················· | ···················· | ···················· | ···················· |
| A | 5692 hCoV-19/France/ARA-487/2020\|EPI_ISL_416747\|2020-03 | ···················· | ···················· | ···················· | ···················· | ···················· | ···················· |
| A | 5693 hCoV-19/France/ARA-508/2020\|EPI_ISL_416748\|2020-03 | ···················· | ···················· | ···················· | ···················· | ···················· | ···················· |
| A | 5694 hCoV-19/France/ARA-532/2020\|EPI_ISL_416749\|2020-03 | ···················· | ···················· | ···················· | ···················· | ···················· | ···················· |
| A | 5695 hCoV-19/France/ARA-683/2020\|EPI_ISL_416750\|2020-03 | ···················· | ···················· | ···················· | ···················· | ···················· | ···················· |
| A | 5696 hCoV-19/France/ARA-651/2020\|EPI_ISL_416751\|2020-03 | ···················· | ···················· | ···················· | ···················· | ···················· | ···················· |
| A | 5697 hCoV-19/France/ARA-650/2020\|EPI_ISL_416752\|2020-03 | ···················· | ···················· | ···················· | ···················· | ···················· | ···················· |
| A | 5698 hCoV-19/France/ARA-06464 /2020\|EPI_ISL_416753\|2020 | ···················· | ···················· | ···················· | ···················· | ···················· | ···················· |
| A | 5699 hCoV-19/France/ARA-06487/2020\|EPI_ISL_416754\|2020- | ···················· | ···················· | ···················· | ···················· | ···················· | ···················· |
| A | 5700 hCoV-19/France/ARA-06531/2020\|EPI_ISL_416756\|2020- | ···················· | ···················· | ···················· | ···················· | ···················· | ···················· |
| A | 5701 hCoV-19/France/ARA-06678/2020\|EPI_ISL_416757\|2020- | ···················· | ···················· | ···················· | ···················· | ···················· | ···················· |
| A | 5702 hCoV-19/France/ARA-0693/2020\|EPI_ISL_416758\|2020-0 | ···················· | ···················· | ···················· | ···················· | ···················· | ···················· |
| A | 5703 hCoV-19/Hangzhou/HZCDC0135/2020\|EPI_ISL_421225\|202 | ···················· | ···················· | ···················· | ···················· | ···················· | ···················· |
| A | 5704 hCoV-19/Hangzhou/HZCDC0119/2020\|EPI_ISL_421226\|202 | ···················· | ···················· | ···················· | ···················· | ···················· | ···················· |
| A | 5705 hCoV-19/Hangzhou/HZCDC0091L/2020\|EPI_ISL_421227\|20 | ···················· | ···················· | ···················· | ···················· | ···················· | ···················· |
| A | 5706 hCoV-19/Hangzhou/HZCDC0091/2020\|EPI_ISL_421228\|202 | ···················· | ···················· | ···················· | ···················· | ···················· | ···················· |
| A | 5707 hCoV-19/Hangzhou/HZCDC0090L/2020\|EPI_ISL_421229\|20 | ···················· | ···················· | ···················· | ···················· | ···················· | ···················· |
| A | 5708 hCoV-19/Hangzhou/HZCDC0090/2020\|EPI_ISL_421230\|202 | ···················· | ···················· | ···················· | ···················· | ···················· | ···················· |
| A | 5709 hCoV-19/Hangzhou/HZCDC0049L/2020\|EPI_ISL_421231\|20 | ···················· | ···················· | ···················· | ···················· | ···················· | ···················· |
| A | 5710 hCoV-19/Hangzhou/HZCDC0048L/2020\|EPI_ISL_421232\|20 | ···················· | ···················· | ···················· | ···················· | ···················· | ···················· |
| A | 5711 hCoV-19/Hangzhou/HZCDC0048/2020\|EPI_ISL_421233\|202 | ···················· | ···················· | ···················· | ···················· | ···················· | ···················· |
| A | 5712 hCoV-19/Hangzhou/HZCDC0025/2020\|EPI_ISL_421234\|202 | ···················· | ···················· | ···················· | ···················· | ···················· | ···················· |
| A | 5713 hCoV-19/Hangzhou/HZCDC0013/2020\|EPI_ISL_421235\|202 | ···················· | ···················· | ···················· | ···················· | ···················· | ···················· |
| A | 5714 hCoV-19/Hangzhou/HZCDC0012/2020\|EPI_ISL_421236\|202 | ···················· | ···················· | ···················· | ···················· | ···················· | ···················· |
| A | 5715 hCoV-19/Jiujiang/JX22/2020\|EPI_ISL_421237\|2020-01- | ···················· | ···················· | ···················· | ···················· | ···················· | ···················· |
| A | 5716 hCoV-19/Nanchang/JX174/2020\|EPI_ISL_421238\|2020-01 | ···················· | ···················· | ···················· | ···················· | ···················· | ···················· |
| A | 5717 hCoV-19/Shangrao/JX105/2020\|EPI_ISL_421239\|2020-01 | ···················· | ···················· | ···················· | ···················· | ···················· | ···················· |
| A | 5718 hCoV-19/Shangrao/JX1948/2020\|EPI_ISL_421240\|2020-0 | ···················· | ···················· | ···················· | ···················· | ···················· | ···················· |
| A | 5719 hCoV-19/Nanchang/JX177/2020\|EPI_ISL_421241\|2020-01 | ···················· | ···················· | ···················· | ···················· | ···················· | ···················· |
| A | 5720 hCoV-19/Ganzhou/JX81/2020\|EPI_ISL_421242\|2020-01-2 | ···················· | ···················· | ···················· | ···················· | ···················· | ···················· |
| A | 5721 hCoV-19/Nanchang/JX14/2020\|EPI_ISL_421243\|2020-01- | ···················· | ···················· | ···················· | ···················· | ···················· | ···················· |
| A | 5722 hCoV-19/Shangrao/JX29/2020\|EPI_ISL_421244\|2020-01- | ···················· | ···················· | ···················· | ···················· | ···················· | ···················· |
| A | 5723 hCoV-19/Nanchang/JX90/2020\|EPI_ISL_421245\|2020-01- | ···················· | ···················· | ···················· | ···················· | ···················· | ···················· |
| A | 5724 hCoV-19/Nanchang/JXN3T4/2020\|EPI_ISL_421246\|2020-0 | ···················· | ···················· | ···················· | ···················· | ···················· | ···················· |
| A | 5725 hCoV-19/Nanchang/JX149/2020\|EPI_ISL_421247\|2020-01 | ···················· | ···················· | ···················· | ···················· | ···················· | ···················· |
| A | 5726 hCoV-19/Nanchang/JX39/2020\|EPI_ISL_421248\|2020-01- | ···················· | ···················· | ···················· | ···················· | ···················· | ···················· |
| A | 5727 hCoV-19/Xinyu/JX122/2020\|EPI_ISL_421249\|2020-01-25 | ···················· | ···················· | ···················· | ···················· | ···················· | ···················· |
| A | 5728 hCoV-19/Shangrao/JX1177/2020\|EPI_ISL_421250\|2020-0 | ···················· | ·A·················· | ···················· | ···················· | ···················· | ···················· |
| A | 5729 hCoV-19/Shangrao/JX1176/2020\|EPI_ISL_421251\|2020-0 | ···················· | ·A·················· | ···················· | ···················· | ···················· | ···················· |
| A | 5730 hCoV-19/Pingxiang/JX5/2020\|EPI_ISL_421252\|2020-01- | ···················· | ···················· | ···················· | ···················· | ···················· | ···················· |
| A | 5731 hCoV-19/Jian/JX129/2020\|EPI_ISL_421253\|2020-01-26 | ···················· | ···················· | ···················· | ···················· | ···················· | ···················· |
| A | 5732 hCoV-19/Shangrao/JX1178/2020\|EPI_ISL_421254\|2020-0 | ···················· | ···················· | ···················· | ···················· | ···················· | ···················· |
| A | 5733 hCoV-19/Jian/JX169/2020\|EPI_ISL_421256\|2020-01-27 | ···················· | ···················· | ···················· | ···················· | ···················· | ···················· |
| A | 5734 hCoV-19/Shangrao/JX1215/2020\|EPI_ISL_421257\|2020-0 | ···················· | ···················· | ···················· | ···················· | ···················· | ···················· |
| A | 5735 hCoV-19/Shangrao/JX1974/2020\|EPI_ISL_421258\|2020-0 | ···················· | ···················· | ···················· | ···················· | ···················· | ···················· |

| | | | | | | | | |
|---|---|---|---|---|---|---|---|---|
| A | 5736 | hCoV-19/Pingxiang/JX151/2020\|EPI_ISL_421259\|2020-0 | .................... | .................... | .................... | .................... | .................... | .................... |
| A | 5737 | hCoV-19/Xinyu/JX124/2020\|EPI_ISL_421260\|2020-01-26 | .................... | .................... | .................... | .................... | .................... | .................... |
| A | 5738 | hCoV-19/Nanchang/JX176/2020\|EPI_ISL_421261\|2020-01 | .................... | .................... | .................... | .................... | .................... | .................... |
| A | 5739 | hCoV-19/Nanchang/JX155/2020\|EPI_ISL_421262\|2020-01 | .................... | .................... | .................... | .................... | .................... | .................... |
| A | 5740 | hCoV-19/USA/WI-UW-29/2020\|EPI_ISL_421287\|2020-03-2 | .................... | .................... | .................... | .................... | .................... | .................... |
| A | 5741 | hCoV-19/USA/WI-UW-30/2020\|EPI_ISL_421288\|2020-03-2 | .................... | .................... | .................... | .................... | .................... | .................... |
| A | 5742 | hCoV-19/USA/WI-UW-31/2020\|EPI_ISL_421289\|2020-03-2 | .................... | .................... | .................... | .................... | .................... | .................... |
| A | 5743 | hCoV-19/USA/NY-PV09301/2020\|EPI_ISL_422556\|2020-03 | .................... | .................... | .................... | .................... | .................... | .................... |
| A | 5744 | hCoV-19/USA/NY-PV09303/2020\|EPI_ISL_422557\|2020-03 | .................... | .................... | .................... | .................... | .................... | .................... |
| A | 5745 | hCoV-19/USA/NY-PV09304/2020\|EPI_ISL_422558\|2020-03 | .................... | .................... | .................... | .................... | .................... | .................... |
| A | 5746 | hCoV-19/USA/NY-PV09305/2020\|EPI_ISL_422559\|2020-03 | .................... | .................... | .................... | .................... | .................... | .................... |
| A | 5747 | hCoV-19/USA/NY-PV09307/2020\|EPI_ISL_422560\|2020-03 | .................... | .................... | .................... | .................... | .................... | .................... |
| A | 5748 | hCoV-19/USA/NY-PV09308/2020\|EPI_ISL_422561\|2020-03 | .................... | .................... | .................... | .................... | .................... | .................... |
| A | 5749 | hCoV-19/USA/NY-PV09309/2020\|EPI_ISL_422562\|2020-03 | .................... | .................... | .................... | .................... | .................... | .................... |
| A | 5750 | hCoV-19/Netherlands/Friesland_10/2020\|EPI_ISL_4225 | .................... | .................... | .................... | .................... | .................... | .................... |
| A | 5751 | hCoV-19/Netherlands/Friesland_3/2020\|EPI_ISL_42256 | .................... | .................... | .................... | .................... | .................... | .................... |
| A | 5752 | hCoV-19/Netherlands/Friesland_4/2020\|EPI_ISL_42256 | .................... | .................... | .................... | .................... | .................... | .................... |
| A | 5753 | hCoV-19/Netherlands/Friesland_6/2020\|EPI_ISL_42257 | .................... | .................... | .................... | .................... | .................... | .................... |
| A | 5754 | hCoV-19/Netherlands/Friesland_7/2020\|EPI_ISL_42257 | .................... | .................... | .................... | .................... | .................... | .................... |
| A | 5755 | hCoV-19/Netherlands/NA_265/2020\|EPI_ISL_422576\|202 | .................... | .................... | .................... | .................... | .................... | .................... |
| A | 5756 | hCoV-19/Netherlands/NA_266/2020\|EPI_ISL_422577\|202 | .................... | .................... | .................... | .................... | .................... | .................... |
| A | 5757 | hCoV-19/Netherlands/NA_267/2020\|EPI_ISL_422578\|202 | .................... | .................... | .................... | .................... | .................... | .................... |
| A | 5758 | hCoV-19/Netherlands/NA_268/2020\|EPI_ISL_422579\|202 | .................... | .................... | .................... | .................... | .................... | .................... |
| A | 5759 | hCoV-19/Netherlands/NA_269/2020\|EPI_ISL_422580\|202 | .................... | .................... | .................... | .................... | .................... | .................... |
| A | 5760 | hCoV-19/Netherlands/NA_270/2020\|EPI_ISL_422581\|202 | .................... | .................... | .................... | .................... | .................... | .................... |
| A | 5761 | hCoV-19/Netherlands/NA_271/2020\|EPI_ISL_422582\|202 | .................... | .................... | .................... | .................... | .................... | .................... |
| A | 5762 | hCoV-19/Netherlands/NA_272/2020\|EPI_ISL_422583\|202 | .................... | .................... | .................... | .................... | .................... | .................... |
| A | 5763 | hCoV-19/Netherlands/NA_274/2020\|EPI_ISL_422585\|202 | .................... | .................... | .................... | .................... | .................... | .................... |
| A | 5764 | hCoV-19/Netherlands/NA_275/2020\|EPI_ISL_422586\|202 | .................... | .................... | .................... | .................... | .................... | .................... |
| A | 5765 | hCoV-19/Netherlands/NA_276/2020\|EPI_ISL_422587\|202 | .................... | .................... | .................... | .................... | .................... | .................... |
| A | 5766 | hCoV-19/Netherlands/NA_277/2020\|EPI_ISL_422588\|202 | .................... | .................... | .................... | .................... | .................... | .................... |
| A | 5767 | hCoV-19/Netherlands/NA_282/2020\|EPI_ISL_422591\|202 | .................... | .................... | .................... | .................... | .................... | .................... |
| A | 5768 | hCoV-19/Netherlands/NA_283/2020\|EPI_ISL_422592\|202 | .................... | .................... | .................... | .................... | .................... | .................... |
| A | 5769 | hCoV-19/Netherlands/NA_284/2020\|EPI_ISL_422593\|202 | .................... | .................... | .................... | .................... | .................... | .................... |
| A | 5770 | hCoV-19/Netherlands/NA_285/2020\|EPI_ISL_422594\|202 | .................... | .................... | .................... | .................... | .................... | .................... |
| A | 5771 | hCoV-19/Netherlands/NA_286/2020\|EPI_ISL_422595\|202 | .................... | .................... | .................... | .................... | .................... | .................... |
| A | 5772 | hCoV-19/Netherlands/NA_287/2020\|EPI_ISL_422596\|202 | .................... | .................... | .................... | .................... | .................... | .................... |
| A | 5773 | hCoV-19/Netherlands/NA_288/2020\|EPI_ISL_422597\|202 | .................... | .................... | .................... | .................... | .................... | .................... |
| A | 5774 | hCoV-19/Netherlands/NA_289/2020\|EPI_ISL_422598\|202 | .................... | ...........A........ | .................... | .................... | .................... | .................... |
| A | 5775 | hCoV-19/Netherlands/NA_290/2020\|EPI_ISL_422599\|202 | .................... | .................... | .................... | .................... | .................... | .................... |
| A | 5776 | hCoV-19/England/20126009804/2020\|EPI_ISL_423887\|20 | .................... | .................... | .................... | .................... | .................... | .................... |
| A | 5777 | hCoV-19/England/20126063104/2020\|EPI_ISL_423903\|20 | .................... | .................... | .................... | .................... | .................... | .................... |
| A | 5778 | hCoV-19/USA/WA-UW-2146/2020\|EPI_ISL_426100\|2020-03 | .................... | .................... | .................... | .................... | .................... | .................... |
| A | 5779 | hCoV-19/USA/WA-UW-2149/2020\|EPI_ISL_426101\|2020-03 | .................... | .................... | .................... | .................... | .................... | .................... |
| A | 5780 | hCoV-19/USA/WA-UW-2164/2020\|EPI_ISL_426102\|2020-03 | .................... | .................... | .................... | .................... | .................... | .................... |
| A | 5781 | hCoV-19/USA/WA-UW-2169/2020\|EPI_ISL_426103\|2020-03 | .................... | .................... | .................... | .................... | .................... | .................... |
| A | 5782 | hCoV-19/USA/WA-UW-2179/2020\|EPI_ISL_426105\|2020-03 | .................... | .................... | .................... | .................... | .................... | .................... |
| A | 5783 | hCoV-19/USA/WA-UW-2180/2020\|EPI_ISL_426106\|2020-03 | .................... | .................... | .................... | .................... | .................... | .................... |
| A | 5784 | hCoV-19/USA/WA-UW-2181/2020\|EPI_ISL_426107\|2020-03 | .................... | .................... | .................... | .................... | .................... | .................... |
| A | 5785 | hCoV-19/USA/WA-UW-2192/2020\|EPI_ISL_426109\|2020-03 | .................... | .................... | .................... | .................... | .................... | .................... |
| A | 5786 | hCoV-19/USA/WA-UW-2203/2020\|EPI_ISL_426111\|2020-03 | .................... | .................... | .................... | .................... | .................... | .................... |
| A | 5787 | hCoV-19/USA/WA-UW-2208/2020\|EPI_ISL_426112\|2020-03 | .................... | .................... | .................... | .................... | .................... | .................... |
| A | 5788 | hCoV-19/USA/WA-UW-2212/2020\|EPI_ISL_426113\|2020-03 | .................... | .................... | .................... | .................... | .................... | .................... |
| A | 5789 | hCoV-19/USA/WA-UW-2213/2020\|EPI_ISL_426114\|2020-03 | .................... | .................... | .................... | .................... | .................... | .................... |
| A | 5790 | hCoV-19/USA/WA-UW-2214/2020\|EPI_ISL_426115\|2020-03 | .................... | .................... | .................... | .................... | .................... | .................... |
| A | 5791 | hCoV-19/USA/WA-UW-2215/2020\|EPI_ISL_426116\|2020-03 | .................... | .................... | .................... | .................... | .................... | .................... |
| A | 5792 | hCoV-19/USA/WA-UW-2218/2020\|EPI_ISL_426117\|2020-03 | .................... | .................... | .................... | .................... | .................... | .................... |
| A | 5793 | hCoV-19/USA/WA-UW-2219/2020\|EPI_ISL_426118\|2020-03 | .................... | .................... | .................... | .................... | .................... | .................... |
| A | 5794 | hCoV-19/USA/WA-UW-2228/2020\|EPI_ISL_426119\|2020-03 | .................... | .................... | .................... | .................... | .................... | .................... |
| A | 5795 | hCoV-19/India/UN-c31/2020\|EPI_ISL_426179\|2020-03-0 | .................... | .................... | .................... | .................... | .................... | .................... |
| A | 5796 | hCoV-19/Qatar/QA13/2020\|EPI_ISL_427408\|2020-03-23 | .................... | .................... | .................... | .................... | .................... | .................... |
| A | 5797 | hCoV-19/Qatar/QA20/2020\|EPI_ISL_427416\|2020-03-23 | .................... | .................... | .................... | .................... | .................... | .................... |
| A | 5798 | hCoV-19/Qatar/QA21/2020\|EPI_ISL_427417\|2020-03-23 | .................... | .................... | .................... | .................... | .................... | .................... |
| A | 5799 | hCoV-19/Qatar/QA22/2020\|EPI_ISL_427418\|2020-03-23 | .................... | .................... | .................... | .................... | .................... | .................... |
| A | 5800 | hCoV-19/USA/WI-UW-120/2020\|EPI_ISL_427427\|2020-04- | .................... | .................... | .................... | .................... | .................... | .................... |
| A | 5801 | hCoV-19/USA/IA-UW-121/2020\|EPI_ISL_427428\|2020-04- | .................... | .................... | .................... | .................... | .................... | .................... |
| A | 5802 | hCoV-19/USA/WI-UW-122/2020\|EPI_ISL_427429\|2020-04- | .................... | .................... | .................... | .................... | .................... | .................... |
| A | 5803 | hCoV-19/USA/IL-UW-123/2020\|EPI_ISL_427430\|2020-04- | .................... | .................... | .................... | .................... | .................... | .................... |
| A | 5804 | hCoV-19/USA/WI-UW-124/2020\|EPI_ISL_427431\|2020-04- | .................... | .................... | .................... | .................... | .................... | .................... |
| A | 5805 | hCoV-19/USA/IL-UW-125/2020\|EPI_ISL_427432\|2020-04- | .................... | .................... | .................... | .................... | .................... | .................... |
| A | 5806 | hCoV-19/USA/IL-UW-126/2020\|EPI_ISL_427433\|2020-04- | .................... | .................... | .................... | .................... | .................... | .................... |
| A | 5807 | hCoV-19/USA/WI-UW-127/2020\|EPI_ISL_427434\|2020-04- | .................... | .................... | .................... | .................... | .................... | .................... |
| A | 5808 | hCoV-19/USA/WI-UW-128/2020\|EPI_ISL_427435\|2020-04- | .................... | .................... | .................... | .................... | .................... | .................... |
| A | 5809 | hCoV-19/USA/WI-UW-129/2020\|EPI_ISL_427436\|2020-04- | .................... | .................... | .................... | .................... | .................... | .................... |
| A | 5810 | hCoV-19/USA/IL-UW-130/2020\|EPI_ISL_427437\|2020-04- | .................... | .................... | .................... | .................... | .................... | .................... |
| A | 5811 | hCoV-19/USA/WI-UW-131/2020\|EPI_ISL_427438\|2020-04- | .................... | .................... | .................... | .................... | .................... | .................... |
| A | 5812 | hCoV-19/USA/WI-UW-132/2020\|EPI_ISL_427439\|2020-04- | .................... | .................... | .................... | .................... | .................... | .................... |
| A | 5813 | hCoV-19/USA/IL-UW-133/2020\|EPI_ISL_427440\|2020-04- | .................... | .................... | .................... | .................... | .................... | .................... |

| | | | | | | | |
|---|---|---|---|---|---|---|---|
| A | 5814 hCoV-19/USA/WI-UW-134/2020\|EPI_ISL_427441\|2020-04- | ···················· | ···················· | ···················· | ···················· | ···················· | ···················· |
| A | 5815 hCoV-19/USA/WI-UW-135/2020\|EPI_ISL_427442\|2020-04- | ···················· | ···················· | ···················· | ···················· | ···················· | ···················· |
| A | 5816 hCoV-19/USA/WI-UW-136/2020\|EPI_ISL_427443\|2020-04- | ···················· | ···················· | ···················· | ···················· | ···················· | ···················· |
| A | 5817 hCoV-19/USA/IL-UW-137/2020\|EPI_ISL_427444\|2020-04- | ···················· | ···················· | ···················· | ···················· | ···················· | ···················· |
| A | 5818 hCoV-19/USA/IL-UW-138/2020\|EPI_ISL_427445\|2020-04- | ···················· | ···················· | ···················· | ···················· | ···················· | ···················· |
| A | 5819 hCoV-19/USA/IL-UW-139/2020\|EPI_ISL_427446\|2020-04- | ···················· | ···················· | ···················· | ···················· | ···················· | ···················· |
| A | 5820 hCoV-19/USA/WI-UW-140/2020\|EPI_ISL_427447\|2020-04- | ···················· | ···················· | ···················· | ···················· | ···················· | ···················· |
| A | 5821 hCoV-19/USA/IL-UW-141/2020\|EPI_ISL_427448\|2020-04- | ···················· | ···················· | ···················· | ···················· | ···················· | ···················· |
| A | 5822 hCoV-19/USA/IL-UW-142/2020\|EPI_ISL_427449\|2020-04- | ···················· | ···················· | ···················· | ···················· | ···················· | ···················· |
| A | 5823 hCoV-19/Spain/MD-H12-2501/2020\|EPI_ISL_428700\|2020 | ···················· | ···················· | ···················· | ···················· | ···················· | ···················· |
| A | 5824 hCoV-19/Spain/MD-H12-2902/2020\|EPI_ISL_428701\|2020 | ···················· | ···················· | ···················· | ···················· | ···················· | ···················· |
| A | 5825 hCoV-19/Spain/MD-H12-3304/2020\|EPI_ISL_428703\|2020 | ···················· | ···················· | ···················· | ···················· | ···················· | ···················· |
| A | 5826 hCoV-19/Spain/MD-H12-3405/2020\|EPI_ISL_428704\|2020 | ···················· | ···················· | ···················· | ···················· | ···················· | ···················· |
| A | 5827 hCoV-19/Spain/MD-H12-3506/2020\|EPI_ISL_428705\|2020 | ···················· | ···················· | ···················· | ···················· | ···················· | ···················· |
| A | 5828 hCoV-19/Spain/MD-H12-3607/2020\|EPI_ISL_428706\|2020 | ···················· | ···················· | ···················· | ···················· | ···················· | ···················· |
| A | 5829 hCoV-19/Spain/MD-H12-3708/2020\|EPI_ISL_428707\|2020 | ···················· | ···················· | ···················· | ···················· | ···················· | ···················· |
| A | 5830 hCoV-19/Spain/MD-H12-3809/2020\|EPI_ISL_428708\|2020 | ···················· | ···················· | ···················· | ···················· | ···················· | ···················· |
| A | 5831 hCoV-19/Spain/MD-H12-3910/2020\|EPI_ISL_428709\|2020 | ···················· | ···················· | ···················· | ···················· | ···················· | ···················· |
| A | 5832 hCoV-19/Spain/MD-H12-4011/2020\|EPI_ISL_428710\|2020 | ···················· | ···················· | ···················· | ···················· | ···················· | ···················· |
| A | 5833 hCoV-19/Spain/MD-H12-4112/2020\|EPI_ISL_428711\|2020 | ···················· | ···················· | ···················· | ···················· | ···················· | ···················· |
| A | 5834 hCoV-19/Turkey/HSGM-4706/2020\|EPI_ISL_428712\|2020- | ···················· | ···················· | ···················· | ···················· | ···················· | ···················· |
| A | 5835 hCoV-19/Turkey/HSGM-5516/2020\|EPI_ISL_428713\|2020- | ···················· | ···················· | ···················· | ···················· | ···················· | ···················· |
| A | 5836 hCoV-19/Turkey/HSGM-5602/2020\|EPI_ISL_428714\|2020- | ···················· | ···················· | ···················· | ···················· | ···················· | ···················· |
| A | 5837 hCoV-19/Turkey/HSGM-5624/2020\|EPI_ISL_428715\|2020- | ···················· | ···················· | ···················· | ···················· | ···················· | ···················· |
| A | 5838 hCoV-19/Turkey/HSGM-5711/2020\|EPI_ISL_428716\|2020- | ···················· | ·A·················· | ···················· | ···················· | ···················· | ··A················· |
| A | 5839 hCoV-19/Turkey/HSGM-5769/2020\|EPI_ISL_428717\|2020- | ···················· | ···················· | ···················· | ···················· | ···················· | ···················· |
| A | 5840 hCoV-19/Turkey/HSGM-5770/2020\|EPI_ISL_428718\|2020- | ···················· | ···················· | ···················· | ···················· | ···················· | ···················· |
| A | 5841 hCoV-19/Turkey/HSGM-7718/2020\|EPI_ISL_428719\|2020- | ···················· | ···················· | ···················· | ···················· | ···················· | ···················· |
| A | 5842 hCoV-19/Turkey/HSGM-8001/2020\|EPI_ISL_428720\|2020- | ···················· | ···················· | ···················· | ···················· | ···················· | ···················· |
| A | 5843 hCoV-19/Turkey/HSGM-8003/2020\|EPI_ISL_428721\|2020- | ···················· | ···················· | ···················· | ···················· | ···················· | ···················· |
| A | 5844 hCoV-19/Turkey/HSGM-8010/2020\|EPI_ISL_428722\|2020- | ···················· | ···················· | ···················· | ···················· | ···················· | ···················· |
| A | 5845 hCoV-19/Turkey/HSGM-8964/2020\|EPI_ISL_428723\|2020- | ···················· | ·A·················· | ···················· | ···················· | ···················· | ··A················· |
| A | 5846 hCoV-19/USA/WI-UW-250/2020\|EPI_ISL_428724\|2020-03- | ···················· | ···················· | ···················· | ···················· | ···················· | ···················· |
| A | 5847 hCoV-19/USA/WI-UW-251/2020\|EPI_ISL_428725\|2020-03- | ···················· | ···················· | ···················· | ···················· | ···················· | ···················· |
| A | 5848 hCoV-19/USA/WI-UW-252/2020\|EPI_ISL_428726\|2020-03- | ···················· | ···················· | ···················· | ···················· | ···················· | ···················· |
| A | 5849 hCoV-19/USA/WI-UW-253/2020\|EPI_ISL_428727\|2020-03- | ···················· | ···················· | ···················· | ···················· | ···················· | ···················· |
| A | 5850 hCoV-19/USA/WI-UW-254/2020\|EPI_ISL_428728\|2020-04- | ···················· | ···················· | ···················· | ···················· | ···················· | ···················· |
| A | 5851 hCoV-19/USA/WI-UW-255/2020\|EPI_ISL_428729\|2020-04- | ···················· | ···················· | ···················· | ···················· | ···················· | ···················· |
| A | 5852 hCoV-19/USA/WI-UW-256/2020\|EPI_ISL_428730\|2020-04- | ···················· | ···················· | ···················· | ···················· | ···················· | ···················· |
| A | 5853 hCoV-19/USA/WI-UW-257/2020\|EPI_ISL_428731\|2020-04- | ···················· | ···················· | ···················· | ···················· | ···················· | ···················· |
| A | 5854 hCoV-19/USA/WI-UW-258/2020\|EPI_ISL_428732\|2020-04- | ···················· | ···················· | ···················· | ···················· | ···················· | ···················· |
| A | 5855 hCoV-19/USA/NY-NYUMC243/2020\|EPI_ISL_428757\|2020-0 | ···················· | ·············A······ | ···················· | ···················· | ···················· | ···················· |
| A | 5856 hCoV-19/USA/NY-NYUMC244/2020\|EPI_ISL_428758\|2020-0 | ···················· | ···················· | ···················· | ···················· | ···················· | ···················· |
| A | 5857 hCoV-19/USA/NY-NYUMC245/2020\|EPI_ISL_428759\|2020-0 | ···················· | ···················· | ···················· | ···················· | ···················· | ···················· |
| A | 5858 hCoV-19/USA/NY-NYUMC246/2020\|EPI_ISL_428760\|2020-0 | ···················· | ···················· | ···················· | ···················· | ···················· | ···················· |
| A | 5859 hCoV-19/USA/NY-NYUMC247/2020\|EPI_ISL_428761\|2020-0 | ···················· | ···················· | ···················· | ···················· | ···················· | ···················· |
| A | 5860 hCoV-19/USA/NY-NYUMC248/2020\|EPI_ISL_428762\|2020-0 | ···················· | ···················· | ···················· | ···················· | ···················· | ···················· |
| A | 5861 hCoV-19/USA/NY-NYUMC249/2020\|EPI_ISL_428763\|2020-0 | ···················· | ···················· | ···················· | ···················· | ···················· | ···················· |
| A | 5862 hCoV-19/USA/NY-NYUMC250/2020\|EPI_ISL_428764\|2020-0 | ···················· | ···················· | ···················· | ···················· | ···················· | ···················· |
| A | 5863 hCoV-19/USA/NY-NYUMC251/2020\|EPI_ISL_428765\|2020-0 | ···················· | ···················· | ···················· | ···················· | ···················· | ···················· |
| A | 5864 hCoV-19/USA/NY-NYUMC252/2020\|EPI_ISL_428766\|2020-0 | ···················· | ···················· | ···················· | ···················· | ···················· | ···················· |
| A | 5865 hCoV-19/USA/NY-NYUMC253/2020\|EPI_ISL_428767\|2020-0 | ···················· | ···················· | ···················· | ···················· | ···················· | ···················· |
| A | 5866 hCoV-19/USA/NY-NYUMC254/2020\|EPI_ISL_428768\|2020-0 | ···················· | ···················· | ···················· | ···················· | ···················· | ···················· |
| A | 5867 hCoV-19/USA/NY-NYUMC255/2020\|EPI_ISL_428769\|2020-0 | ···················· | ···················· | ···················· | ···················· | ···················· | ···················· |
| A | 5868 hCoV-19/USA/NY-NYUMC256/2020\|EPI_ISL_428770\|2020-0 | ···················· | ···················· | ···················· | ···················· | ···················· | ···················· |
| A | 5869 hCoV-19/USA/NY-NYUMC258/2020\|EPI_ISL_428772\|2020-0 | ···················· | ···················· | ···················· | ···················· | ···················· | ···················· |
| A | 5870 hCoV-19/USA/NY-NYUMC259/2020\|EPI_ISL_428773\|2020-0 | ···················· | ···················· | ···················· | ···················· | ···················· | ···················· |
| A | 5871 hCoV-19/USA/NY-NYUMC260/2020\|EPI_ISL_428774\|2020-0 | ···················· | ···················· | ···················· | ···················· | ···················· | ···················· |
| A | 5872 hCoV-19/USA/NY-NYUMC261/2020\|EPI_ISL_428775\|2020-0 | ···················· | ···················· | ···················· | ···················· | ···················· | ···················· |
| A | 5873 hCoV-19/USA/NY-NYUMC262/2020\|EPI_ISL_428776\|2020-0 | ···················· | ···················· | ···················· | ···················· | ···················· | ···················· |
| A | 5874 hCoV-19/USA/NY-NYUMC263/2020\|EPI_ISL_428777\|2020-0 | ···················· | ···················· | ···················· | ···················· | ···················· | ···················· |
| A | 5875 hCoV-19/USA/NY-NYUMC264/2020\|EPI_ISL_428778\|2020-0 | ···················· | ···················· | ···················· | ···················· | ···················· | ···················· |
| A | 5876 hCoV-19/USA/NY-NYUMC265/2020\|EPI_ISL_428779\|2020-0 | ···················· | ···················· | ···················· | ···················· | ···················· | ···················· |
| A | 5877 hCoV-19/Hong Kong/HK20/2020\|EPI_ISL_434560\|2020-01 | ···················· | ···················· | ···················· | ···················· | ···················· | ···················· |
| A | 5878 hCoV-19/Japan/TKYE6968/2020\|EPI_ISL_434561\|2020-02 | ···················· | ···················· | ···················· | ···················· | ···················· | ···················· |
| A | 5879 hCoV-19/Japan/TKYE6947/2020\|EPI_ISL_434562\|2020-03 | ···················· | ···················· | ···················· | ···················· | ···················· | ···················· |
| A | 5880 hCoV-19/Hong Kong/HKU-902a/2020\|EPI_ISL_434563\|202 | ···················· | ···················· | ···················· | ···················· | ···················· | ···················· |
| A | 5881 hCoV-19/Hong Kong/HKU-902b/2020\|EPI_ISL_434564\|202 | ···················· | ···················· | ···················· | ···················· | ···················· | ···················· |
| A | 5882 hCoV-19/Hong Kong/HKU-903a/2020\|EPI_ISL_434565\|202 | ···················· | ···················· | ···················· | ···················· | ···················· | ···················· |
| A | 5883 hCoV-19/Hong Kong/HKU-903b/2020\|EPI_ISL_434566\|202 | ···················· | ···················· | ···················· | ···················· | ···················· | ···················· |
| A | 5884 hCoV-19/Hong Kong/HKU-907a/2020\|EPI_ISL_434567\|202 | ···················· | ···················· | ···················· | ···················· | ···················· | ···················· |
| A | 5885 hCoV-19/Hong Kong/HKU-907b/2020\|EPI_ISL_434568\|202 | ···················· | ···················· | ···················· | ···················· | ···················· | ···················· |
| A | 5886 hCoV-19/Hong Kong/HKU-908a/2020\|EPI_ISL_434569\|202 | ···················· | ···················· | ···················· | ···················· | ···················· | ···················· |
| A | 5887 hCoV-19/Hong Kong/HKU-908b/2020\|EPI_ISL_434570\|202 | ···················· | ···················· | ···················· | ···················· | ···················· | ···················· |
| A | 5888 hCoV-19/Hong Kong/HKU-001a/2020\|EPI_ISL_434571\|202 | ···················· | ···················· | ···················· | ···················· | ···················· | ···················· |
| A | 5889 hCoV-19/USA/CA-CDC-3/2020\|EPI_ISL_408008\|2020-01-2 | ···················· | ·A·················· | ···················· | ···················· | ···················· | ···················· |
| A | 5890 hCoV-19/USA/CA-CDC-4/2020\|EPI_ISL_408009\|2020-01-2 | ···················· | ·A·················· | ···················· | ···················· | ···················· | ···················· |
| A | 5891 hCoV-19/USA/CA-CDC-5/2020\|EPI_ISL_408010\|2020-01-2 | ···················· | ···················· | ···················· | ···················· | ···················· | ···················· |

| | | | | | | | |
|---|---|---|---|---|---|---|---|
| A | 5892 hCoV-19/Nepal/61/2020\|EPI_ISL_410301\|2020-01-13 | ···················· | ···················· | ····················· | ···················· | ···················· | ···················· |
| A | 5893 hCoV-19/Germany/BW-ChVir-1577/2020\|EPI_ISL_412912\| | ···················· | ···················· | ····················· | ···················· | ···················· | ···················· |
| A | 5894 hCoV-19/Brazil/SP-01/2020\|EPI_ISL_412964\|2020-02-2 | ···················· | ···················· | ····················· | ···················· | ···················· | ···················· |
| A | 5895 hCoV-19/Canada/BC_37_0-2/2020\|EPI_ISL_412965\|2020- | ···················· | ···················· | ····················· | ···················· | ···················· | ···················· |
| A | 5896 hCoV-19/Guangzhou/IQTC01/2020\|EPI_ISL_412966\|2020- | ···················· | ···················· | ····················· | ···················· | ···················· | ···················· |
| A | 5897 hCoV-19/Japan/Hu_DP_Kng_19-020/2020\|EPI_ISL_412968 | ···················· | ···················· | ····················· | ···················· | ···················· | ···················· |
| A | 5898 hCoV-19/USA/WA2/2020\|EPI_ISL_412970\|2020-02-24 | ···················· | ···················· | ····················· | ···················· | ···················· | ···················· |
| A | 5899 hCoV-19/Mexico/CMX-InDRE-01/2020\|EPI_ISL_412972\|20 | ···················· | ···················· | ····················· | ···················· | ···················· | ···················· |
| A | 5900 hCoV-19/Italy/LOM-ASST-CDG1/2020\|EPI_ISL_412973\|20 | ···················· | ···················· | ····················· | ···················· | ···················· | ···················· |
| A | 5901 hCoV-19/Italy/LAZ-INMI-SPL1/2020\|EPI_ISL_412974\|20 | ···················· | ···················· | ····················· | ···················· | ···················· | ···················· |
| A | 5902 hCoV-19/Australia/NSW05/2020\|EPI_ISL_412975\|2020-0 | ···················· | ···················· | ····················· | ···················· | ···················· | ···················· |
| A | 5903 hCoV-19/Wuhan/HBCDC-HB-02/2020\|EPI_ISL_412978\|2020 | ···················· | ···················· | ····················· | ···················· | ···················· | ···················· |
| A | 5904 hCoV-19/Wuhan/HBCDC-HB-03/2020\|EPI_ISL_412979\|2020 | ···················· | ···················· | ····················· | ···················· | ···················· | ···················· |
| A | 5905 hCoV-19/Wuhan/HBCDC-HB-04/2020\|EPI_ISL_412980\|2020 | ···················· | ···················· | ····················· | ···················· | ···················· | ···················· |
| A | 5906 hCoV-19/Wuhan/HBCDC-HB-05/2020\|EPI_ISL_412981\|2020 | ···················· | ···················· | ····················· | ···················· | ···················· | ···················· |
| A | 5907 hCoV-19/Wuhan/HBCDC-HB-06/2020\|EPI_ISL_412982\|2020 | ···················· | ···················· | ····················· | ···················· | ···················· | ···················· |
| A | 5908 hCoV-19/Tianmen/HBCDC-HB-07/2020\|EPI_ISL_412983\|20 | ···················· | ···················· | ····················· | ···················· | ···················· | ···················· |
| A | 5909 hCoV-19/Brazil/ES-225/2020\|EPI_ISL_415128\|2020-02- | ···················· | ···················· | ····················· | ···················· | ···················· | ···················· |
| A | 5910 hCoV-19/England/20099038206/2020\|EPI_ISL_415129\|20 | ···················· | ···················· | ····················· | ···················· | ···················· | ···················· |
| A | 5911 hCoV-19/England/20100121006/2020\|EPI_ISL_415140\|20 | ···················· | ···················· | ····················· | ···················· | ···················· | ···················· |
| A | 5912 hCoV-19/England/20100121007/2020\|EPI_ISL_415141\|20 | ···················· | ···················· | ····················· | ···················· | ···················· | ···················· |
| A | 5913 hCoV-19/England/20100122106/2020\|EPI_ISL_415142\|20 | ···················· | ···················· | ····················· | ···················· | ···················· | ···················· |
| A | 5914 hCoV-19/England/20100122107/2020\|EPI_ISL_415143\|20 | ···················· | ···················· | ····················· | ···················· | ···················· | ···················· |
| A | 5915 hCoV-19/England/20102000106/2020\|EPI_ISL_415144\|20 | ···················· | ···················· | ····················· | ···················· | ···················· | ···················· |
| A | 5916 hCoV-19/England/20102000306/2020\|EPI_ISL_415146\|20 | ···················· | ···················· | ····················· | ···················· | ···················· | ···················· |
| A | 5917 hCoV-19/USA/WA-UW84/2020\|EPI_ISL_416440\|2020-03-10 | ···················· | ···················· | ····················· | ···················· | ···················· | ···················· |
| A | 5918 hCoV-19/USA/WA-UW85/2020\|EPI_ISL_416441\|2020-03-10 | ···················· | ···················· | ····················· | ···················· | ···················· | ···················· |
| A | 5919 hCoV-19/USA/WA-UW86/2020\|EPI_ISL_416442\|2020-03-10 | ···················· | ···················· | ····················· | ···················· | ···················· | ···················· |
| A | 5920 hCoV-19/USA/WA-UW87/2020\|EPI_ISL_416443\|2020-03-10 | ···················· | ···················· | ····················· | ···················· | ···················· | ···················· |
| A | 5921 hCoV-19/USA/WA-UW88/2020\|EPI_ISL_416444\|2020-03-10 | ···················· | ···················· | ····················· | ···················· | ···················· | ···················· |
| A | 5922 hCoV-19/USA/WA-UW89/2020\|EPI_ISL_416445\|2020-03-10 | ···················· | ···················· | ····················· | ···················· | ···················· | ···················· |
| A | 5923 hCoV-19/USA/WA-UW90/2020\|EPI_ISL_416446\|2020-03-10 | ···················· | ···················· | ····················· | ···················· | ···················· | ···················· |
| A | 5924 hCoV-19/USA/WA-UW91/2020\|EPI_ISL_416447\|2020-03-10 | ···················· | ···················· | ····················· | ···················· | ···················· | ···················· |
| A | 5925 hCoV-19/USA/WA-UW92/2020\|EPI_ISL_416448\|2020-03-11 | ···················· | ···················· | ····················· | ···················· | ···················· | ···················· |
| A | 5926 hCoV-19/USA/WA-UW93/2020\|EPI_ISL_416449\|2020-03-11 | ···················· | ···················· | ····················· | ···················· | ···················· | ···················· |
| A | 5927 hCoV-19/USA/WA-UW94/2020\|EPI_ISL_416450\|2020-03-11 | ···················· | ···················· | ····················· | ···················· | ···················· | ···················· |
| A | 5928 hCoV-19/USA/WA-UW95/2020\|EPI_ISL_416451\|2020-03-10 | ···················· | ···················· | ····················· | ···················· | ···················· | ···················· |
| A | 5929 hCoV-19/USA/WA-UW96/2020\|EPI_ISL_416452\|2020-03-10 | ···················· | ···················· | ····················· | ···················· | ···················· | ···················· |
| A | 5930 hCoV-19/USA/WA-UW36/2020\|EPI_ISL_416453\|2020-03-07 | ···················· | ···················· | ····················· | ···················· | ···················· | ···················· |
| A | 5931 hCoV-19/USA/WA-UW37/2020\|EPI_ISL_416454\|2020-03-06 | ···················· | ···················· | ····················· | ···················· | ···················· | ···················· |
| A | 5932 hCoV-19/USA/WA-UW38/2020\|EPI_ISL_416455\|2020-03-07 | ···················· | ···················· | ····················· | ···················· | ···················· | ···················· |
| A | 5933 hCoV-19/USA/WA-UW39/2020\|EPI_ISL_416456\|2020-03-06 | ···················· | ···················· | ····················· | ···················· | ···················· | ···················· |
| A | 5934 hCoV-19/USA/CA-MG0987/2020\|EPI_ISL_416457\|2020-03- | ···················· | ···················· | ····················· | ···················· | ···················· | ···················· |
| A | 5935 hCoV-19/Kuwait/KU12/2020\|EPI_ISL_416458\|2020-03-02 | ···················· | ···················· | ····················· | ···················· | ···················· | ···················· |
| A | 5936 hCoV-19/Belgium/DB-03023/2020\|EPI_ISL_416470\|2020- | ···················· | ···················· | ····················· | ···················· | ···················· | ···················· |
| A | 5937 hCoV-19/Belgium/DBD-03024/2020\|EPI_ISL_416471\|2020 | ···················· | ···················· | ····················· | ···················· | ···················· | ···················· |
| A | 5938 hCoV-19/Belgium/UMF-03025/2020\|EPI_ISL_416472\|2020 | ···················· | ···················· | ····················· | ···················· | ···················· | ···················· |
| A | 5939 hCoV-19/Belgium/DBA-03032/2020\|EPI_ISL_416475\|2020 | ···················· | ···················· | ····················· | ···················· | ···················· | ···················· |
| A | 5940 hCoV-19/Georgia/Tb-673/2020\|EPI_ISL_416478\|2020-03 | ···················· | ···················· | ····················· | ···················· | ···················· | ···················· |
| A | 5941 hCoV-19/Georgia/Tb-273/2020\|EPI_ISL_416479\|2020-03 | ···················· | ···················· | ····················· | ···················· | ···················· | ···················· |
| A | 5942 hCoV-19/USA/WA-UW162/2020\|EPI_ISL_416700\|2020-03-1 | ···················· | ···················· | ····················· | ···················· | ···················· | ···················· |
| A | 5943 hCoV-19/USA/WA-UW163/2020\|EPI_ISL_416701\|2020-03-1 | ···················· | ···················· | ····················· | ···················· | ···················· | ···················· |
| A | 5944 hCoV-19/USA/WA-UW164/2020\|EPI_ISL_416702\|2020-03-1 | ···················· | ···················· | ····················· | ···················· | ···················· | ···················· |
| A | 5945 hCoV-19/USA/WA-UW165/2020\|EPI_ISL_416703\|2020-03-1 | ···················· | ···················· | ····················· | ···················· | ···················· | ···················· |
| A | 5946 hCoV-19/USA/WA-UW167/2020\|EPI_ISL_416705\|2020-03-1 | ···················· | ···················· | ····················· | ···················· | ···················· | ···················· |
| A | 5947 hCoV-19/USA/WA-UW168/2020\|EPI_ISL_416706\|2020-03-1 | ···················· | ···················· | ····················· | ···················· | ···················· | ···················· |
| A | 5948 hCoV-19/USA/WA-UW169/2020\|EPI_ISL_416707\|2020-03-1 | ···················· | ···················· | ····················· | ···················· | ···················· | ···················· |
| A | 5949 hCoV-19/USA/WA-UW170/2020\|EPI_ISL_416708\|2020-03-1 | ···················· | ···················· | ····················· | ···················· | ···················· | ···················· |
| A | 5950 hCoV-19/USA/WA-UW171/2020\|EPI_ISL_416709\|2020-03-1 | ···················· | ···················· | ····················· | ···················· | ···················· | ···················· |
| A | 5951 hCoV-19/USA/WA-UW172/2020\|EPI_ISL_416710\|2020-03-1 | ···················· | ···················· | ····················· | ···················· | ···················· | ···················· |
| A | 5952 hCoV-19/USA/WA-UW173/2020\|EPI_ISL_416711\|2020-03-1 | ···················· | ···················· | ····················· | ···················· | ···················· | ···················· |
| A | 5953 hCoV-19/USA/WA-UW174/2020\|EPI_ISL_416712\|2020-03-1 | ···················· | ···················· | ····················· | ···················· | ···················· | ···················· |
| A | 5954 hCoV-19/USA/WA-UW175/2020\|EPI_ISL_416713\|2020-03-1 | ···················· | ···················· | ····················· | ···················· | ···················· | ···················· |
| A | 5955 hCoV-19/USA/WA-UW176/2020\|EPI_ISL_416714\|2020-03-1 | ···················· | ···················· | ····················· | ···················· | ···················· | ···················· |
| A | 5956 hCoV-19/Iceland/112/2020\|EPI_ISL_417770\|2020-03-11 | ···················· | ·A·················· | ····················· | ···················· | ···················· | ···················· |
| A | 5957 hCoV-19/Iceland/131/2020\|EPI_ISL_417771\|2020-03-12 | ···················· | ···················· | ····················· | ···················· | ···················· | ···················· |
| A | 5958 hCoV-19/Iceland/162/2020\|EPI_ISL_417772\|2020-03-14 | ···················· | ···················· | ····················· | ···················· | ···················· | ···················· |
| A | 5959 hCoV-19/Iceland/30/2020\|EPI_ISL_417773\|2020-03-03 | ···················· | ···················· | ····················· | ···················· | ···················· | ···················· |
| A | 5960 hCoV-19/Iceland/175/2020\|EPI_ISL_417774\|2020-03-16 | ···················· | ···················· | ····················· | ···················· | ···················· | ···················· |
| A | 5961 hCoV-19/Iceland/163/2020\|EPI_ISL_417775\|2020-03-14 | ···················· | ···················· | ····················· | ···················· | ···················· | ···················· |
| A | 5962 hCoV-19/Iceland/102/2020\|EPI_ISL_417777\|2020-03-10 | ···················· | ·A·················· | ····················· | ···················· | ···················· | ···················· |
| A | 5963 hCoV-19/Iceland/106/2020\|EPI_ISL_417779\|2020-03-11 | ···················· | ···················· | ····················· | ···················· | ···················· | ···················· |
| A | 5965 hCoV-19/Iceland/123/2020\|EPI_ISL_417783\|2020-03-11 | ···················· | ···················· | ····················· | ···················· | ···················· | ···················· |
| A | 5966 hCoV-19/Iceland/124/2020\|EPI_ISL_417784\|2020-03-12 | ···················· | ···················· | ····················· | ···················· | ···················· | ···················· |
| A | 5967 hCoV-19/Iceland/126/2020\|EPI_ISL_417785\|2020-03-12 | ···················· | ···················· | ····················· | ···················· | ···················· | ···················· |
| A | 5968 hCoV-19/Iceland/128/2020\|EPI_ISL_417786\|2020-03-12 | ···················· | ···················· | ····················· | ···················· | ···················· | ···················· |
| A | 5969 hCoV-19/Iceland/132/2020\|EPI_ISL_417787\|2020-03-12 | ···················· | ···················· | ····················· | ···················· | ···················· | ···················· |
| A | 5970 hCoV-19/Iceland/133/2020\|EPI_ISL_417788\|2020-03-12 | ···················· | ···················· | ····················· | ···················· | ···················· | ···················· |

| | | | | | | | | |
|---|---|---|---|---|---|---|---|---|
| A | 5971 | hCoV-19/Iceland/136/2020\|EPI_ISL_417789\|2020-03-12 | .................... | .................... | ..................... | .................... | .................... | .................... |
| A | 5972 | hCoV-19/USA/WY-WYPHL001/2020\|EPI_ISL_421272\|2020-0 | .................... | .................... | ..................... | .................... | .................... | .................... |
| A | 5973 | hCoV-19/Russia/Moscow_PMVL-1/2020\|EPI_ISL_421275\|2 | .................... | .................... | ..................... | .................... | .................... | .................... |
| A | 5974 | hCoV-19/USA/IN-Lilly-IPB0170-4bc20/2020\|EPI_ISL_42 | .................... | .................... | ..................... | .................... | .................... | .................... |
| A | 5975 | hCoV-19/USA/IN-Lilly-IPB0170-4bc21/2020\|EPI_ISL_42 | .................... | .................... | ..................... | .................... | .................... | .................... |
| A | 5976 | hCoV-19/USA/WI-UW-25/2020\|EPI_ISL_421283\|2020-03-2 | .................... | .................... | ..................... | .................... | .................... | .................... |
| A | 5977 | hCoV-19/USA/WI-UW-26/2020\|EPI_ISL_421284\|2020-03-2 | .................... | .................... | ..................... | .................... | .................... | .................... |
| A | 5978 | hCoV-19/USA/WI-UW-27/2020\|EPI_ISL_421285\|2020-03-2 | .................... | .................... | ..................... | .................... | .................... | .................... |
| A | 5979 | hCoV-19/USA/WI-UW-28/2020\|EPI_ISL_421286\|2020-03-2 | .................... | .................... | ..................... | .................... | .................... | .................... |
| A | 5980 | hCoV-19/USA/WI-UW-32/2020\|EPI_ISL_421290\|2020-03-2 | .................... | .................... | ..................... | .................... | .................... | .................... |
| A | 5981 | hCoV-19/USA/WI-UW-33/2020\|EPI_ISL_421291\|2020-03-2 | .................... | .................... | ..................... | .................... | .................... | .................... |
| A | 5982 | hCoV-19/USA/WI-UW-34/2020\|EPI_ISL_421292\|2020-03-2 | .................... | .................... | ..................... | .................... | .................... | .................... |
| A | 5983 | hCoV-19/USA/WI-UW-35/2020\|EPI_ISL_421293\|2020-03-2 | .................... | .................... | ..................... | .................... | .................... | .................... |
| A | 5984 | hCoV-19/USA/WI-UW-36/2020\|EPI_ISL_421294\|2020-03-2 | .................... | .................... | ..................... | .................... | .................... | .................... |
| A | 5985 | hCoV-19/USA/WI-UW-37/2020\|EPI_ISL_421295\|2020-03-2 | .................... | .................... | ..................... | .................... | .................... | .................... |
| A | 5986 | hCoV-19/USA/WI-UW-38/2020\|EPI_ISL_421296\|2020-03-2 | .................... | .................... | ..................... | .................... | .................... | .................... |
| A | 5987 | hCoV-19/USA/WI-UW-39/2020\|EPI_ISL_421297\|2020-03-2 | .................... | .................... | ..................... | .................... | .................... | .................... |
| A | 5988 | hCoV-19/USA/WI-UW-40/2020\|EPI_ISL_421298\|2020-03-2 | .................... | .................... | ..................... | .................... | .................... | .................... |
| A | 5989 | hCoV-19/USA/WI-UW-41/2020\|EPI_ISL_421299\|2020-03-2 | .................... | .................... | ..................... | .................... | .................... | .................... |
| A | 5991 | hCoV-19/England/20130052804/2020\|EPI_ISL_423632\|20 | .................... | .................... | ..................... | .................... | .................... | .................... |
| A | 5992 | hCoV-19/England/20130061404/2020\|EPI_ISL_423639\|20 | .................... | .................... | ..................... | .................... | .................... | .................... |
| A | 5993 | hCoV-19/USA/IA_6390/2020\|EPI_ISL_424900\|2020-03-07 | .................... | .................... | ..................... | .................... | .................... | .................... |
| A | 5994 | hCoV-19/USA/NJ_3201/2020\|EPI_ISL_424901\|2020-03-04 | .................... | .................... | ..................... | .................... | .................... | .................... |
| A | 5995 | hCoV-19/USA/SC_3520/2020\|EPI_ISL_424902\|2020-03-05 | .................... | .................... | ..................... | .................... | .................... | .................... |
| A | 5996 | hCoV-19/USA/SC_3570/2020\|EPI_ISL_424903\|2020-03-07 | .................... | .................... | ..................... | .................... | .................... | .................... |
| A | 5997 | hCoV-19/USA/SC_3571/2020\|EPI_ISL_424904\|2020-03-07 | .................... | .................... | ..................... | .................... | .................... | .................... |
| A | 5998 | hCoV-19/USA/SC_3572/2020\|EPI_ISL_424905\|2020-03-05 | .................... | .................... | ..................... | .................... | .................... | .................... |
| A | 5999 | hCoV-19/USA/NY_1922/2020\|EPI_ISL_424906\|2020-03-04 | .................... | .................... | ..................... | .................... | .................... | .................... |
| A | 6000 | hCoV-19/USA/VA_6171/2020\|EPI_ISL_424907\|2020-03-06 | .................... | .................... | ..................... | .................... | .................... | .................... |
| A | 6001 | hCoV-19/USA/MA_8932/2020\|EPI_ISL_424908\|2020-03-05 | .................... | .................... | ..................... | .................... | .................... | .................... |
| A | 6002 | hCoV-19/USA/MA_8933/2020\|EPI_ISL_424909\|2020-03-05 | .................... | .................... | ..................... | .................... | .................... | .................... |
| A | 6003 | hCoV-19/USA/MA_9703/2020\|EPI_ISL_424910\|2020-03-05 | .................... | .................... | ..................... | .................... | .................... | .................... |
| A | 6004 | hCoV-19/USA/MA_9704/2020\|EPI_ISL_424911\|2020-03-04 | .................... | .................... | ..................... | .................... | .................... | .................... |
| A | 6005 | hCoV-19/USA/MA_9889/2020\|EPI_ISL_424912\|2020-03-04 | .................... | .................... | ..................... | .................... | .................... | .................... |
| A | 6006 | hCoV-19/USA/MA_1355/2020\|EPI_ISL_424913\|2020-03-05 | .................... | .................... | ..................... | .................... | .................... | .................... |
| A | 6007 | hCoV-19/USA/MA_2652/2020\|EPI_ISL_424914\|2020-03-06 | .................... | .................... | ..................... | .................... | .................... | .................... |
| A | 6008 | hCoV-19/USA/MA_3614/2020\|EPI_ISL_424915\|2020-03-06 | .................... | .................... | ..................... | .................... | .................... | .................... |
| A | 6009 | hCoV-19/USA/MA_3616/2020\|EPI_ISL_424916\|2020-03-06 | .................... | .................... | ..................... | .................... | .................... | .................... |
| A | 6010 | hCoV-19/USA/MA_3623/2020\|EPI_ISL_424917\|2020-03-06 | .................... | .................... | ..................... | .................... | .................... | .................... |
| A | 6011 | hCoV-19/USA/MA_3626/2020\|EPI_ISL_424918\|2020-03-06 | .................... | .................... | ..................... | .................... | .................... | .................... |
| A | 6012 | hCoV-19/USA/MA_3642/2020\|EPI_ISL_424919\|2020-03-07 | .................... | .................... | ..................... | .................... | .................... | .................... |
| A | 6013 | hCoV-19/USA/MA_3653/2020\|EPI_ISL_424920\|2020-03-06 | .................... | .................... | ..................... | .................... | .................... | .................... |
| A | 6014 | hCoV-19/USA/NY-NYUMC122/2020\|EPI_ISL_424929\|2020-0 | .................... | .................... | ..................... | .................... | .................... | .................... |
| A | 6015 | hCoV-19/USA/NY-NYUMC123/2020\|EPI_ISL_424930\|2020-0 | .................... | .................... | ..................... | .................... | .................... | .................... |
| A | 6016 | hCoV-19/USA/NY-NYUMC124/2020\|EPI_ISL_424931\|2020-0 | .................... | .................... | ..................... | .................... | .................... | .................... |
| A | 6017 | hCoV-19/USA/NY-NYUMC125/2020\|EPI_ISL_424932\|2020-0 | .................... | .................... | ..................... | .................... | .................... | .................... |
| A | 6018 | hCoV-19/USA/NY-NYUMC126/2020\|EPI_ISL_424933\|2020-0 | .................... | .................... | ..................... | .................... | .................... | .................... |
| A | 6019 | hCoV-19/USA/NY-NYUMC127/2020\|EPI_ISL_424934\|2020-0 | .................... | .................... | ..................... | .................... | .................... | .................... |
| A | 6020 | hCoV-19/USA/NY-NYUMC128/2020\|EPI_ISL_424935\|2020-0 | .................... | .................... | ..................... | .................... | .................... | .................... |
| A | 6021 | hCoV-19/USA/NY-NYUMC129/2020\|EPI_ISL_424936\|2020-0 | .................... | .................... | ..................... | .................... | .................... | .................... |
| A | 6022 | hCoV-19/USA/NY-NYUMC130/2020\|EPI_ISL_424937\|2020-0 | .................... | .................... | ..................... | .................... | .................... | .................... |
| A | 6023 | hCoV-19/USA/NY-NYUMC131/2020\|EPI_ISL_424938\|2020-0 | .................... | .................... | ..................... | .................... | .................... | .................... |
| A | 6024 | hCoV-19/USA/NY-NYUMC132/2020\|EPI_ISL_424939\|2020-0 | .................... | .................... | ..................... | .................... | .................... | .................... |
| A | 6025 | hCoV-19/USA/NY-NYUMC133/2020\|EPI_ISL_424940\|2020-0 | .................... | .................... | ..................... | .................... | .................... | .................... |
| A | 6026 | hCoV-19/USA/NY-NYUMC134/2020\|EPI_ISL_424941\|2020-0 | .................... | .................... | ..................... | .................... | .................... | .................... |
| A | 6027 | hCoV-19/USA/NY-NYUMC135/2020\|EPI_ISL_424942\|2020-0 | .................... | .................... | ..................... | .................... | .................... | .................... |
| A | 6028 | hCoV-19/USA/NY-NYUMC136/2020\|EPI_ISL_424943\|2020-0 | .................... | .................... | ..................... | .................... | .................... | .................... |
| A | 6029 | hCoV-19/USA/NY-NYUMC137/2020\|EPI_ISL_424944\|2020-0 | .................... | .................... | ..................... | .................... | .................... | .................... |
| A | 6030 | hCoV-19/USA/NY-NYUMC138/2020\|EPI_ISL_424945\|2020-0 | .................... | .................... | ..................... | .................... | .................... | .................... |
| A | 6031 | hCoV-19/USA/NY-NYUMC139/2020\|EPI_ISL_424946\|2020-0 | .................... | .................... | ..................... | .................... | .................... | .................... |
| A | 6032 | hCoV-19/USA/NY-NYUMC140/2020\|EPI_ISL_424947\|2020-0 | .................... | .................... | ..................... | .................... | .................... | .................... |
| A | 6033 | hCoV-19/USA/NY-NYUMC141/2020\|EPI_ISL_424948\|2020-0 | .................... | .................... | ..................... | .................... | .................... | .................... |
| A | 6034 | hCoV-19/USA/NY-NYUMC142/2020\|EPI_ISL_424949\|2020-0 | .................... | .................... | ..................... | .................... | .................... | .................... |
| A | 6035 | hCoV-19/USA/NY-NYUMC143/2020\|EPI_ISL_424950\|2020-0 | .................... | .................... | ..................... | .................... | .................... | .................... |
| A | 6036 | hCoV-19/USA/NY-NYUMC144/2020\|EPI_ISL_424951\|2020-0 | .................... | .................... | ..................... | .................... | .................... | .................... |
| A | 6037 | hCoV-19/USA/NY-NYUMC145/2020\|EPI_ISL_424952\|2020-0 | .................... | .................... | ..................... | .................... | .................... | .................... |
| A | 6038 | hCoV-19/USA/NY-NYUMC146/2020\|EPI_ISL_424953\|2020-0 | .................... | .................... | ..................... | .................... | .................... | .................... |
| A | 6039 | hCoV-19/USA/NY-NYUMC147/2020\|EPI_ISL_424954\|2020-0 | .................... | .................... | ..................... | .................... | .................... | .................... |
| A | 6040 | hCoV-19/USA/NY-NYUMC148/2020\|EPI_ISL_424955\|2020-0 | .................... | .................... | ..................... | .................... | .................... | .................... |
| A | 6041 | hCoV-19/USA/NY-NYUMC149/2020\|EPI_ISL_424956\|2020-0 | .................... | .................... | ..................... | .................... | .................... | .................... |
| A | 6042 | hCoV-19/USA/NY-NYUMC150/2020\|EPI_ISL_424957\|2020-0 | .................... | .................... | ..................... | .................... | .................... | .................... |
| A | 6043 | hCoV-19/USA/NY-NYUMC151/2020\|EPI_ISL_424958\|2020-0 | .................... | .................... | ..................... | .................... | .................... | .................... |
| A | 6044 | hCoV-19/USA/NY-NYUMC152/2020\|EPI_ISL_424959\|2020-0 | .................... | .................... | ..................... | .................... | .................... | .................... |
| A | 6045 | hCoV-19/USA/NY-NYUMC153/2020\|EPI_ISL_424960\|2020-0 | .................... | .................... | ..................... | .................... | .................... | .................... |
| A | 6046 | hCoV-19/USA/NY-NYUMC154/2020\|EPI_ISL_424961\|2020-0 | .................... | .................... | ..................... | .................... | .................... | .................... |
| A | 6047 | hCoV-19/USA/NY-NYUMC155/2020\|EPI_ISL_424962\|2020-0 | .................... | .................... | ..................... | .................... | .................... | .................... |
| A | 6048 | hCoV-19/USA/NY-NYUMC156/2020\|EPI_ISL_424963\|2020-0 | .................... | .................... | ..................... | .................... | .................... | .................... |
| A | 6049 | hCoV-19/USA/NY-NYUMC157/2020\|EPI_ISL_424964\|2020-0 | .................... | .................... | ..................... | .................... | .................... | .................... |

```
A    6050 hCoV-19/USA/NY-NYUMC158/2020|EPI_ISL_424965|2020-0    ····················  ····················  ····················  ····················  ····················  ····················
A    6051 hCoV-19/USA/NY-NYUMC159/2020|EPI_ISL_424966|2020-0    ····················  ····················  ····················  ····················  ····················  ····················
A    6052 hCoV-19/USA/NY-NYUMC160/2020|EPI_ISL_424967|2020-0    ····················  ····················  ····················  ····················  ····················  ····················
A    6053 hCoV-19/USA/NY-NYUMC161/2020|EPI_ISL_424968|2020-0    ····················  ····················  ····················  ····················  ····················  ····················
A    6054 hCoV-19/Taiwan/CGMH-CGU-13/2020|EPI_ISL_424969|202    ····················  ····················  ····················  ····················  ····················  ····················
A    6055 hCoV-19/Taiwan/CGMH-CGU-14/2020|EPI_ISL_424970|202    ····················  ····················  ····················  ····················  ····················  ····················
A    6056 hCoV-19/Taiwan/CGMH-CGU-15/2020|EPI_ISL_424971|202    ····················  ····················  ····················  ····················  ····················  ····················
A    6057 hCoV-19/Taiwan/CGMH-CGU-16/2020|EPI_ISL_424972|202    ····················  ····················  ····················  ····················  ····················  ····················
A    6058 hCoV-19/Taiwan/CGMH-CGU-17/2020|EPI_ISL_424973|202    ····················  ····················  ····················  ····················  ····················  ····················
A    6059 hCoV-19/Taiwan/CGMH-CGU-18/2020|EPI_ISL_424974|202    ····················  ····················  ····················  ····················  ····················  ····················
A    6060 hCoV-19/Taiwan/CGMH-CGU-19/2020|EPI_ISL_424975|202    ····················  ····················  ····················  ····················  ····················  ····················
A    6062 hCoV-19/Australia/VIC851/2020|EPI_ISL_427111|2020-    ····················  ····················  ····················  ····················  ····················  ····················
A    6063 hCoV-19/Australia/VIC853/2020|EPI_ISL_427113|2020-    ····················  ····················  ····················  ····················  ····················  ····················
A    6064 hCoV-19/Australia/VIC856/2020|EPI_ISL_427115|2020-    ····················  ····················  ····················  ····················  ····················  ····················
A    6065 hCoV-19/Australia/VIC857/2020|EPI_ISL_427116|2020-    ····················  ····················  ····················  ····················  ····················  ····················
A    6066 hCoV-19/Australia/VIC858/2020|EPI_ISL_427117|2020-    ····················  ····················  ····················  ····················  ····················  ····················
A    6067 hCoV-19/Australia/VIC860/2020|EPI_ISL_427118|2020-    ····················  ····················  ····················  ····················  ····················  ····················
A    6068 hCoV-19/Australia/VIC862/2020|EPI_ISL_427119|2020-    ····················  ····················  ····················  ····················  ····················  ····················
A    6069 hCoV-19/Australia/VIC863/2020|EPI_ISL_427120|2020-    ····················  ····················  ····················  ····················  ····················  ····················
A    6070 hCoV-19/Australia/VIC864/2020|EPI_ISL_427121|2020-    ····················  ····················  ····················  ····················  ····················  ····················
A    6071 hCoV-19/Australia/VIC867/2020|EPI_ISL_427122|2020-    ····················  ····················  ····················  ····················  ····················  ····················
A    6072 hCoV-19/Australia/VIC868/2020|EPI_ISL_427123|2020-    ····················  ····················  ····················  ····················  ····················  ····················
A    6073 hCoV-19/Australia/VIC869/2020|EPI_ISL_427124|2020-    ····················  ····················  ····················  ····················  ····················  ····················
A    6075 hCoV-19/Australia/VIC871/2020|EPI_ISL_427126|2020-    ····················  ····················  ····················  ····················  ····················  ····················
A    6076 hCoV-19/Australia/VIC873/2020|EPI_ISL_427128|2020-    ····················  ····················  ····················  ····················  ····················  ····················
A    6077 hCoV-19/Australia/VIC874/2020|EPI_ISL_427129|2020-    ····················  ····················  ····················  ····················  ····················  ····················
A    6078 hCoV-19/USA/ID-UW-4266/2020|EPI_ISL_427170|2020-03    ····················  ····················  ····················  ····················  ····················  ····················
A    6079 hCoV-19/USA/ID-UW-4462/2020|EPI_ISL_427171|2020-03    ····················  ····················  ····················  ····················  ····················  ····················
A    6080 hCoV-19/USA/ID-UW-4486/2020|EPI_ISL_427172|2020-03    ····················  ····················  ····················  ····················  ····················  ····················
A    6081 hCoV-19/USA/OR-UW-4300/2020|EPI_ISL_427173|2020-03    ····················  ····················  ····················  ····················  ····················  ····················
A    6082 hCoV-19/USA/WA-UW-3809/2020|EPI_ISL_427174|2020-03    ····················  ····················  ····················  ····················  ····················  ····················
A    6083 hCoV-19/USA/WA-UW-3810/2020|EPI_ISL_427175|2020-03    ····················  ····················  ····················  ····················  ····················  ····················
A    6084 hCoV-19/USA/WA-UW-3814/2020|EPI_ISL_427177|2020-03    ····················  ····················  ····················  ····················  ····················  ····················
A    6085 hCoV-19/USA/WA-UW-3815/2020|EPI_ISL_427178|2020-03    ····················  ····················  ····················  ····················  ····················  ····················
A    6086 hCoV-19/USA/WA-UW-3816/2020|EPI_ISL_427179|2020-03    ····················  ····················  ····················  ····················  ····················  ····················
A    6087 hCoV-19/USA/WA-UW-3819/2020|EPI_ISL_427180|2020-03    ····················  ····················  ····················  ····················  ····················  ····················
A    6088 hCoV-19/USA/WA-UW-3826/2020|EPI_ISL_427181|2020-03    ····················  ····················  ····················  ····················  ····················  ····················
A    6089 hCoV-19/USA/WA-UW-3827/2020|EPI_ISL_427182|2020-03    ····················  ····················  ····················  ····················  ····················  ····················
A    6090 hCoV-19/USA/WA-UW-3828/2020|EPI_ISL_427183|2020-03    ····················  ····················  ····················  ····················  ····················  ····················
A    6091 hCoV-19/USA/WA-UW-3829/2020|EPI_ISL_427184|2020-03    ····················  ····················  ····················  ····················  ····················  ····················
A    6092 hCoV-19/USA/WA-UW-3832/2020|EPI_ISL_427185|2020-03    ····················  ····················  ····················  ····················  ····················  ····················
A    6093 hCoV-19/USA/WA-UW-3834/2020|EPI_ISL_427187|2020-03    ····················  ····················  ····················  ····················  ····················  ····················
A    6094 hCoV-19/USA/WA-UW-3838/2020|EPI_ISL_427188|2020-03    ····················  ····················  ····················  ····················  ····················  ····················
A    6095 hCoV-19/USA/WA-UW-3839/2020|EPI_ISL_427189|2020-03    ····················  ····················  ····················  ····················  ····················  ····················
A    6096 hCoV-19/USA/WA-UW-3841/2020|EPI_ISL_427190|2020-03    ····················  ····················  ····················  ····················  ····················  ····················
A    6097 hCoV-19/USA/WA-UW-3843/2020|EPI_ISL_427191|2020-03    ····················  ····················  ····················  ····················  ····················  ····················
A    6098 hCoV-19/USA/WA-UW-3844/2020|EPI_ISL_427192|2020-03    ····················  ····················  ····················  ····················  ····················  ····················
A    6099 hCoV-19/USA/WA-UW-3850/2020|EPI_ISL_427193|2020-03    ····················  ····················  ····················  ····················  ····················  ····················
A    6100 hCoV-19/USA/WA-UW-3855/2020|EPI_ISL_427194|2020-03    ····················  ····················  ····················  ····················  ····················  ····················
A    6101 hCoV-19/USA/WA-UW-3858/2020|EPI_ISL_427195|2020-03    ····················  ····················  ····················  ····················  ····················  ····················
A    6102 hCoV-19/USA/WA-UW-3862/2020|EPI_ISL_427196|2020-03    ····················  ····················  ····················  ····················  ····················  ····················
A    6103 hCoV-19/USA/WA-UW-3868/2020|EPI_ISL_427197|2020-03    ····················  ····················  ····················  ····················  ····················  ····················
A    6104 hCoV-19/USA/WA-UW-3871/2020|EPI_ISL_427198|2020-03    ····················  ····················  ····················  ····················  ····················  ····················
A    6105 hCoV-19/USA/WA-UW-3880/2020|EPI_ISL_427199|2020-03    ····················  ····················  ····················  ····················  ····················  ····················
A    6106 hCoV-19/Guangdong/20SF1107/2020|EPI_ISL_428441|202    ····················  ····················  ····················  ····················  ····················  ····················
A    6107 hCoV-19/Guangdong/20SF1136/2020|EPI_ISL_428442|202    ····················  ····················  ····················  ····················  ····················  ····················
A    6108 hCoV-19/Guangdong/20SF115/2020|EPI_ISL_428443|2020    ····················  ····················  ····················  ····················  ····················  ····················
A    6109 hCoV-19/Guangdong/20SF1152/2020|EPI_ISL_428444|202    ····················  ·A··················  ····················  ····················  ····················  ····················
A    6110 hCoV-19/Guangdong/20SF1153/2020|EPI_ISL_428445|202    ····················  ·A··················  ····················  ····················  ····················  ····················
A    6111 hCoV-19/Guangdong/20SF1159/2020|EPI_ISL_428446|202    ····················  ····················  ····················  ····················  ····················  ····················
A    6112 hCoV-19/Guangdong/20SF117/2020|EPI_ISL_428447|2020    ····················  ····················  ····················  ····················  ····················  ····················
A    6113 hCoV-19/Guangdong/20SF118/2020|EPI_ISL_428448|2020    ····················  ····················  ····················  ····················  ····················  ····················
A    6114 hCoV-19/Guangdong/20SF123/2020|EPI_ISL_428449|2020    ····················  ····················  ····················  ····················  ····················  ····················
A    6115 hCoV-19/Guangdong/20SF190/2020|EPI_ISL_428452|2020    ····················  ·A··················  ····················  ····················  ····················  ····················
A    6116 hCoV-19/Guangdong/20SF198/2020|EPI_ISL_428454|2020    ····················  ····················  ····················  ····················  ····················  ····················
A    6117 hCoV-19/Guangdong/20SF200/2020|EPI_ISL_428455|2020    ····················  ····················  ····················  ····················  ····················  ····················
A    6118 hCoV-19/Guangdong/20SF243/2020|EPI_ISL_428456|2020    ····················  ····················  ····················  ····················  ····················  ····················
A    6119 hCoV-19/Guangdong/20SF254/2020|EPI_ISL_428457|2020    ····················  ····················  ····················  ····················  ····················  ····················
A    6120 hCoV-19/Guangdong/20SF2546/2020|EPI_ISL_428458|202    ····················  ····················  ····················  ····················  ····················  ····················
A    6121 hCoV-19/Luxembourg/LNS4530810/2020|EPI_ISL_429770|    ····················  ····················  ····················  ····················  ····················  ····················
A    6122 hCoV-19/Luxembourg/LNS4107430/2020|EPI_ISL_429772|    ····················  ····················  ····················  ····················  ····················  ····················
A    6123 hCoV-19/Luxembourg/LNS3257811/2020|EPI_ISL_429773|    ····················  ····················  ····················  ····················  ····················  ····················
A    6124 hCoV-19/Luxembourg/LNS5611124/2020|EPI_ISL_429774|    ····················  ····················  ····················  ····················  ····················  ····················
A    6125 hCoV-19/Luxembourg/LNS7299024/2020|EPI_ISL_429775|    ····················  ····················  ····················  ····················  ····················  ····················
A    6126 hCoV-19/Luxembourg/LNS8726389/2020|EPI_ISL_429776|    ····················  ····················  ····················  ····················  ····················  ····················
A    6127 hCoV-19/Luxembourg/LNS0994857/2020|EPI_ISL_429777|    ····················  ····················  ····················  ····················  ····················  ····················
A    6128 hCoV-19/Luxembourg/LNS2580723/2020|EPI_ISL_429778|    ····················  ····················  ····················  ····················  ····················  ····················
A    6129 hCoV-19/Luxembourg/LNS7357097/2020|EPI_ISL_429779|    ····················  ····················  ····················  ····················  ····················  ····················
```

| | | | | | | | |
|---|---|---|---|---|---|---|---|
| A | 6130 hCoV-19/Luxembourg/LNS5680075/2020\|EPI_ISL_429780\| | .................... | .................... | .................... | .................... | .................... | .................... |
| A | 6131 hCoV-19/Luxembourg/LNS0709551/2020\|EPI_ISL_429781\| | .................... | .................... | .................... | .................... | .................... | .................... |
| A | 6132 hCoV-19/Luxembourg/LNS3169999/2020\|EPI_ISL_429782\| | .................... | .................... | .................... | .................... | .................... | .................... |
| A | 6133 hCoV-19/Luxembourg/LNS5677421/2020\|EPI_ISL_429784\| | .................... | .................... | .................... | .................... | .................... | .................... |
| A | 6134 hCoV-19/Luxembourg/LNS4818581/2020\|EPI_ISL_429786\| | .................... | .................... | .................... | .................... | .................... | .................... |
| A | 6135 hCoV-19/Luxembourg/LNS3283768/2020\|EPI_ISL_429787\| | .................... | .................... | .................... | .................... | .................... | .................... |
| A | 6136 hCoV-19/Guangzhou/IQTC03/2020\|EPI_ISL_529149\|2020- | .................... | .................... | .................... | .................... | .................... | .................... |
| A | 6137 hCoV-19/Guangzhou/IQTC04/2020\|EPI_ISL_529150\|2020- | .................... | .................... | .................... | .................... | .................... | .................... |
| A | 6138 hCoV-19/USA/CT-UW420/2020\|EPI_ISL_529155\|2020-03-2 | .................... | .................... | .................... | .................... | .................... | .................... |
| A | 6139 hCoV-19/USA/CT-UW423/2020\|EPI_ISL_529156\|2020-03-2 | .................... | .................... | .................... | .................... | .................... | .................... |
| A | 6140 hCoV-19/USA/CT-UW427/2020\|EPI_ISL_529157\|2020-03-2 | .................... | .................... | .................... | .................... | .................... | .................... |
| A | 6141 hCoV-19/USA/CT-UW430/2020\|EPI_ISL_529159\|2020-03-2 | .................... | .................... | .................... | .................... | .................... | .................... |
| A | 6142 hCoV-19/USA/ID-UW-2173/2020\|EPI_ISL_529160\|2020-03 | .................... | .................... | .................... | .................... | .................... | .................... |
| A | 6143 hCoV-19/USA/ID-UW-2195/2020\|EPI_ISL_529161\|2020-03 | .................... | .................... | .................... | .................... | .................... | .................... |
| A | 6144 hCoV-19/USA/OR-UW409/2020\|EPI_ISL_529170\|2020-03-2 | .................... | .................... | .................... | .................... | .................... | .................... |
| A | 6145 hCoV-19/Brazil/BA-312/2020\|EPI_ISL_415105\|2020-03- | .................... | .................... | .................... | .................... | .................... | .................... |
| A | 6146 hCoV-19/England/20099079106/2020\|EPI_ISL_415130\|20 | .................... | .................... | .................... | .................... | .................... | .................... |
| A | 6147 hCoV-19/England/20100004706/2020\|EPI_ISL_415133\|20 | .................... | .................... | .................... | .................... | .................... | .................... |
| A | 6148 hCoV-19/England/20100004806/2020\|EPI_ISL_415134\|20 | .................... | .................... | .................... | .................... | .................... | .................... |
| A | 6150 hCoV-19/England/20102000206/2020\|EPI_ISL_415145\|20 | .................... | .................... | .................... | .................... | .................... | ...........Y........ |
| A | 6151 hCoV-19/England/20102000506/2020\|EPI_ISL_415147\|20 | .................... | .................... | .................... | .................... | .................... | .................... |
| A | 6154 hCoV-19/USA/NY2-PV08100/2020\|EPI_ISL_415151\|2020-0 | .................... | .................... | .................... | .................... | .................... | .................... |
| A | 6155 hCoV-19/Panama/328677/2020\|EPI_ISL_415152\|2020-03- | .................... | .................... | .................... | .................... | .................... | .................... |
| A | 6156 hCoV-19/Belgium/VLM-03011/2020\|EPI_ISL_415153\|2020 | .................... | .................... | .................... | .................... | .................... | .................... |
| A | 6157 hCoV-19/Belgium/BM-03012/2020\|EPI_ISL_415154\|2020- | .................... | .................... | .................... | .................... | .................... | .................... |
| A | 6158 hCoV-19/Belgium/VAG-03013/2020\|EPI_ISL_415155\|2020 | .................... | .................... | .................... | .................... | .................... | .................... |
| A | 6159 hCoV-19/Belgium/SH-03014/2020\|EPI_ISL_415156\|2020- | .................... | .................... | .................... | .................... | .................... | .................... |
| A | 6160 hCoV-19/Belgium/BC-03016/2020\|EPI_ISL_415157\|2020- | .................... | .................... | .................... | .................... | .................... | .................... |
| A | 6161 hCoV-19/Belgium/QKJ-03015/2020\|EPI_ISL_415158\|2020 | .................... | .................... | .................... | .................... | .................... | .................... |
| A | 6162 hCoV-19/Belgium/BA-02291/2020\|EPI_ISL_415159\|2020- | .................... | .................... | .................... | .................... | .................... | .................... |
| A | 6163 hCoV-19/Shanghai/SH0112/2020\|EPI_ISL_416400\|2020-0 | .................... | .................... | .................... | .................... | .................... | .................... |
| A | 6164 hCoV-19/Shanghai/SH0114/2020\|EPI_ISL_416401\|2020-0 | .................... | .................... | .................... | .................... | .................... | .................... |
| A | 6165 hCoV-19/Shanghai/SH0115/2020\|EPI_ISL_416402\|2020-0 | .................... | .................... | .................... | .................... | .................... | .................... |
| A | 6166 hCoV-19/Shanghai/SH0117/2020\|EPI_ISL_416403\|2020-0 | .................... | .................... | .................... | .................... | .................... | .................... |
| A | 6167 hCoV-19/Shanghai/SH0119/2020\|EPI_ISL_416404\|2020-0 | .................... | .................... | .................... | .................... | .................... | .................... |
| A | 6168 hCoV-19/Shanghai/SH0121/2020\|EPI_ISL_416405\|2020-0 | .................... | .................... | .................... | .................... | .................... | .................... |
| A | 6169 hCoV-19/Shanghai/SH0125/2020\|EPI_ISL_416406\|2020-0 | .................... | .................... | .................... | .................... | .................... | .................... |
| A | 6170 hCoV-19/Shanghai/SH0126/2020\|EPI_ISL_416407\|2020-0 | .................... | .................... | .................... | .................... | .................... | .................... |
| A | 6171 hCoV-19/Shanghai/SH0128/2020\|EPI_ISL_416409\|2020-0 | .................... | .................... | .................... | .................... | .................... | .................... |
| A | 6172 hCoV-19/Australia/VIC02/2020\|EPI_ISL_416410\|2020-0 | .................... | .................... | .................... | .................... | .................... | .................... |
| A | 6173 hCoV-19/Australia/VIC03/2020\|EPI_ISL_416411\|2020-0 | .................... | .................... | .................... | .................... | .................... | .................... |
| A | 6174 hCoV-19/Australia/VIC04/2020\|EPI_ISL_416412\|2020-0 | .................... | .................... | .................... | .................... | .................... | .................... |
| A | 6175 hCoV-19/Australia/VIC05/2020\|EPI_ISL_416413\|2020-0 | .................... | .................... | .................... | .................... | .................... | .................... |
| A | 6176 hCoV-19/Australia/VIC07/2020\|EPI_ISL_416415\|2020-0 | .................... | .................... | .................... | .................... | .................... | .................... |
| A | 6177 hCoV-19/Hungary/mbl1/2020\|EPI_ISL_416426\|2020-03-1 | .................... | .................... | .................... | .................... | .................... | .................... |
| A | 6178 hCoV-19/Vietnam/38142/2020\|EPI_ISL_416427\|2020-01- | .................... | .................... | .................... | .................... | .................... | .................... |
| A | 6179 hCoV-19/Vietnam/39607/2020\|EPI_ISL_416428\|2020-03- | .................... | .................... | .................... | .................... | .................... | .................... |
| A | 6180 hCoV-19/Vietnam/CM99/2020\|EPI_ISL_416429\|2020-02-1 | .................... | .................... | .................... | .................... | .................... | .................... |
| A | 6181 hCoV-19/Vietnam/CM295/2020\|EPI_ISL_416430\|2020-03- | .................... | .................... | .................... | .................... | .................... | .................... |
| A | 6182 hCoV-19/Vietnam/CM296/2020\|EPI_ISL_416431\|2020-03- | .................... | .................... | .................... | .................... | .................... | .................... |
| A | 6183 hCoV-19/Saudi Arabia/KAIMRC-Alghoribi/2020\|EPI_ISL | .................... | .................... | .................... | .................... | .................... | .................... |
| A | 6184 hCoV-19/USA/WA-UW77/2020\|EPI_ISL_416433\|2020-03-10 | .................... | .................... | .................... | .................... | .................... | .................... |
| A | 6185 hCoV-19/USA/WA-UW78/2020\|EPI_ISL_416434\|2020-03-10 | .................... | .................... | .................... | .................... | .................... | .................... |
| A | 6186 hCoV-19/USA/WA-UW79/2020\|EPI_ISL_416435\|2020-03-10 | .................... | .................... | .................... | .................... | .................... | .................... |
| A | 6187 hCoV-19/USA/WA-UW80/2020\|EPI_ISL_416436\|2020-03-10 | .................... | .................... | .................... | .................... | .................... | .................... |
| A | 6188 hCoV-19/USA/WA-UW81/2020\|EPI_ISL_416437\|2020-03-10 | .................... | .................... | .................... | .................... | .................... | .................... |
| A | 6189 hCoV-19/USA/WA-UW82/2020\|EPI_ISL_416438\|2020-03-10 | .................... | .................... | .................... | .................... | .................... | .................... |
| A | 6190 hCoV-19/USA/WA-UW83/2020\|EPI_ISL_416439\|2020-03-10 | .................... | .................... | .................... | .................... | .................... | .................... |
| A | 6191 hCoV-19/USA/WA-S5/2020\|EPI_ISL_416460\|2020-02-29 | .................... | .................... | .................... | .................... | .................... | .................... |
| A | 6192 hCoV-19/USA/WA-S6/2020\|EPI_ISL_416461\|2020-02-29 | .................... | .................... | .................... | .................... | .................... | .................... |
| A | 6193 hCoV-19/USA/WA-S7/2020\|EPI_ISL_416462\|2020-02-24 | .................... | .................... | .................... | .................... | .................... | .................... |
| A | 6194 hCoV-19/USA/WA-S10/2020\|EPI_ISL_416465\|2020-02-29 | .................... | .................... | .................... | .................... | .................... | .................... |
| A | 6195 hCoV-19/USA/WA-S11/2020\|EPI_ISL_416466\|2020-03-03 | .................... | .................... | .................... | .................... | .................... | .................... |
| A | 6196 hCoV-19/Belgium/MTR-03021/2020\|EPI_ISL_416467\|2020 | .................... | .................... | .................... | .................... | .................... | .................... |
| A | 6197 hCoV-19/Belgium/GMH-03022/2020\|EPI_ISL_416468\|2020 | .................... | .................... | .................... | .................... | .................... | .................... |
| A | 6198 hCoV-19/Belgium/SN-03031/2020\|EPI_ISL_416469\|2020- | .................... | .................... | .................... | .................... | .................... | .................... |
| A | 6199 hCoV-19/Belgium/MTR-03026/2020\|EPI_ISL_416476\|2020 | .................... | .................... | .................... | .................... | .................... | .................... |
| A | 6200 hCoV-19/Georgia/Tb-390/2020\|EPI_ISL_416477\|2020-03 | .................... | .................... | .................... | .................... | .................... | .................... |
| A | 6201 hCoV-19/Georgia/Tb-537/2020\|EPI_ISL_416480\|2020-03 | .................... | .................... | .................... | .................... | .................... | .................... |
| A | 6202 hCoV-19/Georgia/Tb-712/2020\|EPI_ISL_416481\|2020-03 | .................... | .................... | .................... | .................... | .................... | .................... |
| A | 6203 hCoV-19/Georgia/Tb/2020\|EPI_ISL_416482\|2020-03-13 | .................... | .................... | .................... | .................... | .................... | .................... |
| A | 6204 hCoV-19/Spain/VC-FISABIO-5/2020\|EPI_ISL_416484\|202 | .................... | .................... | .................... | .................... | .................... | .................... |
| A | 6205 hCoV-19/Spain/VC-FISABIO-6/2020\|EPI_ISL_416485\|202 | .................... | .................... | .................... | .................... | .................... | .................... |
| A | 6206 hCoV-19/Spain/VC-FISABIO-7/2020\|EPI_ISL_416486\|202 | .................... | .................... | .................... | .................... | .................... | .................... |
| A | 6207 hCoV-19/Spain/VC-FISABIO-8/2020\|EPI_ISL_416487\|202 | .................... | .................... | .................... | .................... | .................... | .................... |
| A | 6208 hCoV-19/Poland/PL_P1/2020\|EPI_ISL_416488\|2020-03-0 | .................... | .................... | .................... | .................... | .................... | .................... |
| A | 6209 hCoV-19/USA/WI-UW-02/2020\|EPI_ISL_416489\|2020-03-1 | .................... | .................... | .................... | .................... | .................... | .................... |
| A | 6210 hCoV-19/USA/WI-UW-04/2020\|EPI_ISL_416491\|2020-03-1 | .................... | .................... | .................... | .................... | .................... | .................... |

| | | | | | | | | | |
|---|---|---|---|---|---|---|---|---|---|
| A | 6211 hCoV-19/USA/WI-UW-05/2020\|EPI_ISL_416492\|2020-03-1 | ························· | ························· | ························· | ························· | ························· | ························· | | |
| A | 6212 hCoV-19/France/HDF-2196/2020\|EPI_ISL_416493\|2020-0 | ························· | ························· | ························· | ························· | ························· | ························· | | |
| A | 6213 hCoV-19/France/NOR-N2223/2020\|EPI_ISL_416494\|2020- | ························· | ························· | ························· | ························· | ························· | ························· | | |
| A | 6214 hCoV-19/France/HDF-2234/2020\|EPI_ISL_416495\|2020-0 | ························· | ························· | ························· | ························· | ························· | ························· | | |
| A | 6215 hCoV-19/France/HDF-2237/2020\|EPI_ISL_416496\|2020-0 | ························· | ························· | ························· | ························· | ························· | ························· | | |
| A | 6216 hCoV-19/France/HDF-2239/2020\|EPI_ISL_416497\|2020-0 | ························· | ························· | ························· | ························· | ························· | ························· | | |
| A | 6217 hCoV-19/France/IDF-2256/2020\|EPI_ISL_416498\|2020-0 | ························· | ························· | ························· | ························· | ························· | ························· | | |
| A | 6218 hCoV-19/France/IDF-2278/2020\|EPI_ISL_416499\|2020-0 | ························· | ························· | ························· | ························· | ························· | ························· | | |
| A | 6219 hCoV-19/Iceland/173/2020\|EPI_ISL_417700\|2020-03-15 | ························· | ························· | ························· | ························· | ························· | ························· | | |
| A | 6222 hCoV-19/Iceland/179/2020\|EPI_ISL_417703\|2020-03-16 | ························· | ························· | ························· | ························· | ························· | ························· | | |
| A | 6223 hCoV-19/Iceland/92/2020\|EPI_ISL_417704\|2020-03-10 | ························· | ························· | ························· | ························· | ························· | ·····················N·NN | 1 | 1 |
| A | 6224 hCoV-19/Iceland/65/2020\|EPI_ISL_417705\|2020-03-07 | ························· | ························· | ························· | ························· | ························· | ························· | | |
| A | 6225 hCoV-19/Iceland/199/2020\|EPI_ISL_417706\|2020-03-16 | ························· | ························· | ························· | ························· | ························· | ························· | | |
| A | 6226 hCoV-19/Iceland/189/2020\|EPI_ISL_417709\|2020-03-16 | ························· | ························· | ························· | ························· | ························· | ························· | | |
| A | 6227 hCoV-19/Iceland/150/2020\|EPI_ISL_417711\|2020-03-13 | ························· | ························· | ························· | ························· | ························· | ························· | | |
| A | 6228 hCoV-19/Iceland/172/2020\|EPI_ISL_417712\|2020-03-15 | ························· | ························· | ························· | ························· | ························· | ························· | | |
| A | 6229 hCoV-19/Iceland/130/2020\|EPI_ISL_417713\|2020-03-12 | ························· | ························· | ························· | ························· | ························· | ························· | | |
| A | 6230 hCoV-19/Iceland/129/2020\|EPI_ISL_417714\|2020-03-12 | ························· | ························· | ························· | ························· | ························· | ························· | | |
| A | 6231 hCoV-19/Iceland/45/2020\|EPI_ISL_417715\|2020-03-04 | ························· | ························· | ························· | ························· | ························· | ························· | | |
| A | 6232 hCoV-19/Iceland/174/2020\|EPI_ISL_417716\|2020-03-16 | ························· | ························· | ························· | ························· | ························· | ························· | | |
| A | 6233 hCoV-19/Iceland/195/2020\|EPI_ISL_417717\|2020-03-16 | ························· | ························· | ························· | ························· | ························· | ························· | | |
| A | 6234 hCoV-19/Iceland/49/2020\|EPI_ISL_417718\|2020-03-05 | ························· | ························· | ························· | ························· | ························· | ························· | | |
| A | 6235 hCoV-19/Iceland/138/2020\|EPI_ISL_417720\|2020-03-12 | ························· | ························· | ························· | ························· | ························· | ························· | | |
| A | 6236 hCoV-19/Iceland/27/2020\|EPI_ISL_417721\|2020-03-03 | ························· | ························· | ························· | ························· | ························· | ························· | | |
| A | 6237 hCoV-19/Iceland/110/2020\|EPI_ISL_417722\|2020-03-11 | ························· | ························· | ························· | ························· | ························· | ························· | | |
| A | 6238 hCoV-19/Iceland/182/2020\|EPI_ISL_417724\|2020-03-16 | ························· | ························· | ························· | ························· | ························· | ························· | | |
| A | 6239 hCoV-19/Iceland/161/2020\|EPI_ISL_417725\|2020-03-14 | ························· | ························· | ························· | ························· | ························· | ························· | | |
| A | 6241 hCoV-19/Iceland/127/2020\|EPI_ISL_417727\|2020-03-12 | ························· | ························· | ························· | ························· | ························· | ························· | | |
| A | 6242 hCoV-19/Iceland/70/2020\|EPI_ISL_417728\|2020-03-08 | ························· | ························· | ························· | ························· | ························· | ························· | | |
| A | 6243 hCoV-19/Iceland/93/2020\|EPI_ISL_417730\|2020-03-10 | ························· | ························· | ························· | ························· | ························· | ························· | | |
| A | 6244 hCoV-19/Iceland/61/2020\|EPI_ISL_417731\|2020-03-06 | ························· | ························· | ························· | ························· | ························· | ························· | | |
| A | 6245 hCoV-19/Iceland/94/2020\|EPI_ISL_417732\|2020-03-10 | ························· | ························· | ························· | ························· | ························· | ························· | | |
| A | 6246 hCoV-19/Iceland/207/2020\|EPI_ISL_417733\|2020-03-16 | ························· | ························· | ························· | ························· | ························· | ························· | | |
| A | 6247 hCoV-19/Iceland/22/2020\|EPI_ISL_417734\|2020-03-02 | ························· | ························· | ························· | ························· | ························· | ························· | | |
| A | 6248 hCoV-19/Iceland/134/2020\|EPI_ISL_417735\|2020-03-12 | ························· | ························· | ························· | ························· | ························· | ························· | | |
| A | 6249 hCoV-19/Iceland/14/2020\|EPI_ISL_417736\|2020-03-01 | ························· | ························· | ························· | ························· | ························· | ························· | | |
| A | 6250 hCoV-19/Iceland/155/2020\|EPI_ISL_417737\|2020-03-15 | ························· | ························· | ························· | ························· | ························· | ························· | | |
| A | 6251 hCoV-19/Iceland/89/2020\|EPI_ISL_417738\|2020-03-10 | ························· | ························· | ························· | ························· | ························· | ························· | | |
| A | 6252 hCoV-19/Iceland/5/2020\|EPI_ISL_417739\|2020-03-13 | ························· | ························· | ························· | ························· | ························· | ························· | | |
| A | 6253 hCoV-19/Iceland/7/2020\|EPI_ISL_417740\|2020-03-15 | ························· | ························· | ························· | ························· | ························· | ························· | | |
| A | 6254 hCoV-19/Iceland/145/2020\|EPI_ISL_417741\|2020-03-13 | ························· | ························· | ························· | ························· | ························· | ························· | | |
| A | 6255 hCoV-19/Iceland/197/2020\|EPI_ISL_417742\|2020-03-16 | ························· | ························· | ························· | ························· | ························· | ························· | | |
| A | 6256 hCoV-19/Iceland/200/2020\|EPI_ISL_417743\|2020-03-16 | ························· | ························· | ························· | ························· | ························· | ························· | | |
| A | 6257 hCoV-19/Iceland/137/2020\|EPI_ISL_417744\|2020-03-13 | ························· | ························· | ························· | ························· | ························· | ························· | | |
| A | 6258 hCoV-19/Iceland/152/2020\|EPI_ISL_417745\|2020-03-13 | ························· | ························· | ························· | ························· | ························· | ························· | | |
| A | 6259 hCoV-19/Iceland/183/2020\|EPI_ISL_417746\|2020-03-16 | ························· | ························· | ························· | ························· | ························· | ························· | | |
| A | 6261 hCoV-19/Iceland/109/2020\|EPI_ISL_417748\|2020-03-11 | ························· | ························· | ························· | ························· | ························· | ························· | | |
| A | 6262 hCoV-19/Iceland/69/2020\|EPI_ISL_417749\|2020-03-08 | ························· | ························· | ························· | ························· | ························· | ························· | | |
| A | 6263 hCoV-19/Iceland/202/2020\|EPI_ISL_417750\|2020-03-16 | ························· | ························· | ························· | ························· | ························· | ························· | | |
| A | 6264 hCoV-19/Iceland/213/2020\|EPI_ISL_417752\|2020-03-16 | ························· | ························· | ························· | ························· | ························· | ························· | | |
| A | 6265 hCoV-19/Iceland/180/2020\|EPI_ISL_417753\|2020-03-16 | ························· | ························· | ························· | ························· | ························· | ························· | | |
| A | 6266 hCoV-19/Iceland/226/2020\|EPI_ISL_417754\|2020-03-16 | ························· | ························· | ························· | ························· | ························· | ························· | | |
| A | 6267 hCoV-19/Iceland/169/2020\|EPI_ISL_417755\|2020-03-13 | ························· | ························· | ························· | ························· | ························· | ························· | | |
| A | 6268 hCoV-19/Iceland/62/2020\|EPI_ISL_417757\|2020-03-06 | ························· | ························· | ························· | ························· | ························· | ························· | | |
| A | 6269 hCoV-19/Iceland/164/2020\|EPI_ISL_417758\|2020-03-14 | ························· | ························· | ························· | ························· | ························· | ························· | | |
| A | 6270 hCoV-19/Iceland/166/2020\|EPI_ISL_417759\|2020-03-13 | ························· | ························· | ························· | ························· | ························· | ························· | | |
| A | 6271 hCoV-19/Iceland/84/2020\|EPI_ISL_417761\|2020-03-10 | ························· | ························· | ························· | ························· | ························· | ························· | | |
| A | 6272 hCoV-19/Iceland/191/2020\|EPI_ISL_417762\|2020-03-16 | ························· | ························· | ························· | ························· | ························· | ························· | | |
| A | 6273 hCoV-19/Iceland/211/2020\|EPI_ISL_417763\|2020-03-16 | ························· | ························· | ························· | ························· | ························· | ························· | | |
| A | 6274 hCoV-19/Iceland/209/2020\|EPI_ISL_417764\|2020-03-16 | ························· | ························· | ························· | ························· | ························· | ························· | | |
| A | 6275 hCoV-19/Iceland/13/2020\|EPI_ISL_417765\|2020-02-27 | ························· | ························· | ························· | ························· | ························· | ························· | | |
| A | 6276 hCoV-19/Iceland/208/2020\|EPI_ISL_417766\|2020-03-16 | ························· | ·A······················· | ························· | ························· | ························· | ························· | | |
| A | 6278 hCoV-19/Iceland/139/2020\|EPI_ISL_417790\|2020-03-12 | ························· | ························· | ························· | ························· | ························· | ························· | | |
| A | 6279 hCoV-19/Iceland/140/2020\|EPI_ISL_417791\|2020-03-12 | ························· | ························· | ························· | ························· | ························· | ························· | | |
| A | 6280 hCoV-19/Iceland/141/2020\|EPI_ISL_417792\|2020-03-13 | ························· | ························· | ························· | ························· | ························· | ························· | | |
| A | 6281 hCoV-19/Iceland/142/2020\|EPI_ISL_417793\|2020-03-13 | ························· | ························· | ························· | ························· | ························· | ························· | | |
| A | 6282 hCoV-19/Iceland/144/2020\|EPI_ISL_417795\|2020-03-13 | ························· | ························· | ························· | ························· | ························· | ························· | | |
| A | 6283 hCoV-19/Iceland/146/2020\|EPI_ISL_417796\|2020-03-13 | ························· | ························· | ························· | ························· | ························· | ························· | | |
| A | 6284 hCoV-19/Iceland/147/2020\|EPI_ISL_417797\|2020-03-13 | ························· | ························· | ························· | ························· | ························· | ························· | | |
| A | 6285 hCoV-19/Iceland/148/2020\|EPI_ISL_417798\|2020-03-13 | ························· | ·A······················· | ························· | ························· | ························· | ························· | | |
| A | 6286 hCoV-19/Iceland/149/2020\|EPI_ISL_417799\|2020-03-13 | ························· | ························· | ························· | ························· | ························· | ························· | | |
| A | 6287 hCoV-19/Australia/VIC836/2020\|EPI_ISL_427100\|2020- | ························· | ························· | ························· | ························· | ························· | ························· | | |
| A | 6288 hCoV-19/Australia/VIC837/2020\|EPI_ISL_427101\|2020- | ························· | ························· | ························· | ························· | ························· | ························· | | |
| A | 6289 hCoV-19/Australia/VIC842/2020\|EPI_ISL_427102\|2020- | ························· | ························· | ························· | ························· | ························· | ························· | | |
| A | 6290 hCoV-19/Australia/VIC843/2020\|EPI_ISL_427103\|2020- | ························· | ························· | ························· | ························· | ························· | ························· | | |
| A | 6291 hCoV-19/Australia/VIC845/2020\|EPI_ISL_427105\|2020- | ························· | ························· | ························· | ························· | ························· | ························· | | |
| A | 6292 hCoV-19/Australia/VIC846/2020\|EPI_ISL_427106\|2020- | ························· | ························· | ························· | ························· | ························· | ························· | | |
| A | 6293 hCoV-19/Australia/VIC847/2020\|EPI_ISL_427107\|2020- | ························· | ························· | ························· | ························· | ························· | ························· | | |

| | | | | | | | | |
|---|---|---|---|---|---|---|---|---|
| A | 6294 | hCoV-19/Australia/VIC848/2020\|EPI_ISL_427108\|2020- | .................... | .................... | .................... | .................... | .................... | .................... |
| A | 6295 | hCoV-19/Australia/VIC849/2020\|EPI_ISL_427109\|2020- | .................... | .................... | .................... | .................... | .................... | .................... |
| A | 6296 | hCoV-19/Australia/VIC875/2020\|EPI_ISL_427130\|2020- | .................... | .................... | .................... | .................... | .................... | .................... |
| A | 6297 | hCoV-19/Australia/VIC876/2020\|EPI_ISL_427131\|2020- | .................... | .................... | .................... | .................... | .................... | .................... |
| A | 6298 | hCoV-19/Australia/VIC878/2020\|EPI_ISL_427133\|2020- | .................... | .................... | .................... | .................... | .................... | .................... |
| A | 6299 | hCoV-19/Australia/VIC879/2020\|EPI_ISL_427134\|2020- | .................... | .................... | .................... | .................... | .................... | .................... |
| A | 6300 | hCoV-19/Australia/VIC881/2020\|EPI_ISL_427135\|2020- | .................... | .................... | .................... | .................... | .................... | .................... |
| A | 6301 | hCoV-19/Australia/VIC883/2020\|EPI_ISL_427136\|2020- | .................... | .................... | .................... | .................... | .................... | .................... |
| A | 6303 | hCoV-19/Australia/VIC886/2020\|EPI_ISL_427139\|2020- | .................... | .................... | .................... | .................... | .................... | .................... |
| A | 6304 | hCoV-19/Australia/VIC887/2020\|EPI_ISL_427140\|2020- | .................... | .................... | .................... | .................... | .................... | .................... |
| A | 6306 | hCoV-19/Australia/VIC893/2020\|EPI_ISL_427145\|2020- | .................... | .................... | .................... | .................... | .................... | .................... |
| A | 6308 | hCoV-19/Australia/VIC895/2020\|EPI_ISL_427147\|2020- | .................... | .................... | .................... | .................... | .................... | .................... |
| A | 6309 | hCoV-19/Australia/VIC898/2020\|EPI_ISL_427148\|2020- | .................... | .................... | .................... | .................... | .................... | .................... |
| A | 6310 | hCoV-19/Australia/VIC899/2020\|EPI_ISL_427149\|2020- | .................... | .................... | .................... | .................... | .................... | .................... |
| A | 6315 | hCoV-19/Australia/VIC931/2020\|EPI_ISL_427159\|2020- | .................... | .................... | .................... | .................... | .................... | .................... |
| A | 6316 | hCoV-19/Australia/VIC932/2020\|EPI_ISL_427160\|2020- | .................... | .................... | .................... | .................... | .................... | .................... |
| A | 6317 | hCoV-19/USA/CT-UW-3845/2020\|EPI_ISL_427161\|2020-03 | .................... | .................... | .................... | .................... | .................... | .................... |
| A | 6318 | hCoV-19/USA/CT-UW-4238/2020\|EPI_ISL_427162\|2020-03 | .................... | .................... | .................... | .................... | .................... | .................... |
| A | 6319 | hCoV-19/USA/CT-UW-4244/2020\|EPI_ISL_427163\|2020-03 | .................... | .................... | .................... | .................... | .................... | .................... |
| A | 6320 | hCoV-19/USA/CT-UW-4251/2020\|EPI_ISL_427164\|2020-03 | .................... | .................... | .................... | .................... | .................... | .................... |
| A | 6322 | hCoV-19/USA/ID-UW-3888/2020\|EPI_ISL_427166\|2020-03 | .................... | .................... | .................... | .................... | .................... | .................... |
| A | 6323 | hCoV-19/USA/ID-UW-3892/2020\|EPI_ISL_427167\|2020-03 | .................... | .................... | .................... | .................... | .................... | .................... |
| A | 6324 | hCoV-19/USA/ID-UW-4239/2020\|EPI_ISL_427169\|2020-03 | .................... | .................... | .................... | .................... | .................... | .................... |
| A | 6325 | hCoV-19/Guangdong/20SF273/2020\|EPI_ISL_428460\|2020 | .................... | .................... | .................... | .................... | .................... | .................... |
| A | 6327 | hCoV-19/Guangdong/20SF4047/2020\|EPI_ISL_428462\|202 | .................... | .................... | .................... | .................... | .................... | .................... |
| A | 6328 | hCoV-19/Guangdong/20SF4051/2020\|EPI_ISL_428463\|202 | .................... | .................... | .................... | .................... | .................... | .................... |
| A | 6329 | hCoV-19/Guangdong/20SF602/2020\|EPI_ISL_428464\|2020 | .................... | .................... | .................... | .................... | .................... | .................... |
| A | 6330 | hCoV-19/Guangdong/20SF616/2020\|EPI_ISL_428465\|2020 | .................... | .................... | .................... | .................... | .................... | .................... |
| A | 6331 | hCoV-19/Guangdong/20SF629/2020\|EPI_ISL_428466\|2020 | .................... | .................... | .................... | .................... | .................... | .................... |
| A | 6332 | hCoV-19/Guangdong/20SF630/2020\|EPI_ISL_428467\|2020 | .................... | .................... | .................... | .................... | .................... | .................... |
| A | 6333 | hCoV-19/Guangdong/20SF632/2020\|EPI_ISL_428468\|2020 | .................... | .................... | .................... | .................... | .................... | .................... |
| A | 6334 | hCoV-19/Guangdong/20SF665/2020\|EPI_ISL_428469\|2020 | .................... | .................... | .................... | .................... | .................... | .................... |
| A | 6335 | hCoV-19/Guangdong/20SF684/2020\|EPI_ISL_428470\|2020 | .................... | .................... | .................... | .................... | .................... | .................... |
| A | 6336 | hCoV-19/Guangdong/20SF685/2020\|EPI_ISL_428471\|2020 | .................... | .................... | .................... | .................... | .................... | .................... |
| A | 6337 | hCoV-19/Guangdong/20SF753/2020\|EPI_ISL_428472\|2020 | .................... | .................... | .................... | .................... | .................... | .................... |
| A | 6338 | hCoV-19/Guangdong/20SF758/2020\|EPI_ISL_428473\|2020 | .................... | .................... | .................... | .................... | .................... | .................... |
| A | 6339 | hCoV-19/Guangdong/20SF808/2020\|EPI_ISL_428474\|2020 | .................... | .................... | .................... | .................... | .................... | .................... |
| A | 6340 | hCoV-19/Guangdong/20SF812/2020\|EPI_ISL_428475\|2020 | .................... | .................... | .................... | .................... | .................... | .................... |
| A | 6341 | hCoV-19/Guangdong/20SF813/2020\|EPI_ISL_428476\|2020 | .................... | .................... | .................... | .................... | .................... | .................... |
| A | 6342 | hCoV-19/Guangdong/20SF822/2020\|EPI_ISL_428477\|2020 | .................... | .................... | .................... | .................... | .................... | .................... |
| A | 6343 | hCoV-19/Guangdong/20SF840/2020\|EPI_ISL_428478\|2020 | .................... | .................... | .................... | .................... | .................... | .................... |
| A | 6344 | hCoV-19/India/KA-nimh-0113/2020\|EPI_ISL_428479\|202 | .................... | .................... | .................... | .................... | .................... | .................... |
| A | 6346 | hCoV-19/India/KA-nimh-0130/2020\|EPI_ISL_428481\|202 | .................... | .................... | .................... | .................... | .................... | .................... |
| A | 6347 | hCoV-19/India/KA-nimh-0182/2020\|EPI_ISL_428482\|202 | .................... | .................... | .................... | .................... | .................... | .................... |
| A | 6348 | hCoV-19/India/KA-nimh-0318/2020\|EPI_ISL_428483\|202 | .................... | .................... | .................... | .................... | .................... | .................... |
| A | 6349 | hCoV-19/India/KA-nimh-0351/2020\|EPI_ISL_428484\|202 | .................... | .................... | .................... | .................... | .................... | .................... |
| A | 6350 | hCoV-19/India/KA-nimh-0996/2020\|EPI_ISL_428486\|202 | .................... | .................... | .................... | .................... | .................... | .................... |
| A | 6351 | hCoV-19/India/KA-nimh-1071/2020\|EPI_ISL_428487\|202 | .................... | .................... | .................... | .................... | .................... | .................... |
| A | 6352 | hCoV-19/Taiwan/8/2020\|EPI_ISL_428488\|2020-01-31 | .................... | .................... | .................... | .................... | .................... | .................... |
| A | 6353 | hCoV-19/Taiwan/5/2020\|EPI_ISL_428489\|2020-01-31 | .................... | .................... | .................... | .................... | .................... | .................... |
| A | 6354 | hCoV-19/Taiwan/78/2020\|EPI_ISL_428490\|2020-03-16 | .................... | .................... | .................... | .................... | .................... | .................... |
| A | 6355 | hCoV-19/Taiwan/58/2020\|EPI_ISL_428491\|2020-03-15 | .................... | .................... | .................... | .................... | .................... | .................... |
| A | 6357 | hCoV-19/Croatia/AU-S10/2020\|EPI_ISL_429705\|2020-03 | .................... | .................... | .................... | .................... | .................... | .................... |
| A | 6358 | hCoV-19/Luxembourg/LNS3652733/2020\|EPI_ISL_429706\| | .................... | .................... | .................... | .................... | .................... | .................... |
| A | 6359 | hCoV-19/Luxembourg/LNS4641661/2020\|EPI_ISL_429707\| | .................... | .................... | .................... | .................... | .................... | .................... |
| A | 6360 | hCoV-19/Luxembourg/LNS5041225/2020\|EPI_ISL_429710\| | .................... | .................... | .................... | .................... | .................... | .................... |
| A | 6361 | hCoV-19/Luxembourg/LNS5519604/2020\|EPI_ISL_429711\| | .................... | .................... | .................... | .................... | .................... | .................... |
| A | 6362 | hCoV-19/Luxembourg/LNS9831617/2020\|EPI_ISL_429713\| | .................... | .................... | .................... | .................... | .................... | .................... |
| A | 6363 | hCoV-19/Luxembourg/LNS7469301/2020\|EPI_ISL_429715\| | .................... | .................... | .................... | .................... | .................... | .................... |
| A | 6364 | hCoV-19/Luxembourg/LNS7032690/2020\|EPI_ISL_429716\| | .................... | .................... | .................... | .................... | .................... | .................... |
| A | 6365 | hCoV-19/Luxembourg/LNS1995247/2020\|EPI_ISL_429718\| | .................... | .................... | .................... | .................... | .................... | .................... |
| A | 6366 | hCoV-19/Luxembourg/LNS5277361/2020\|EPI_ISL_429723\| | .................... | .................... | .................... | .................... | .................... | .................... |
| A | 6367 | hCoV-19/Luxembourg/LNS3212178/2020\|EPI_ISL_429728\| | .................... | .................... | .................... | .................... | .................... | .................... |
| A | 6368 | hCoV-19/Luxembourg/LNS0481305/2020\|EPI_ISL_429729\| | .................... | .................... | .................... | .................... | .................... | .................... |
| A | 6369 | hCoV-19/Luxembourg/LNS7438855/2020\|EPI_ISL_429730\| | .................... | .................... | .................... | .................... | .................... | .................... |
| A | 6370 | hCoV-19/Luxembourg/LNS7580831/2020\|EPI_ISL_429732\| | .................... | .................... | .................... | .................... | .................... | .................... |
| A | 6371 | hCoV-19/Luxembourg/LNS4423730/2020\|EPI_ISL_429733\| | .................... | .................... | .................... | .................... | .................... | .................... |
| A | 6372 | hCoV-19/Luxembourg/LNS6783622/2020\|EPI_ISL_429734\| | .................... | .................... | .................... | .................... | .................... | .................... |
| A | 6373 | hCoV-19/Luxembourg/LNS5358505/2020\|EPI_ISL_429735\| | .................... | .................... | .................... | .................... | .................... | .................... |
| A | 6374 | hCoV-19/Luxembourg/LNS7640155/2020\|EPI_ISL_429736\| | .................... | .................... | .................... | .................... | .................... | .................... |
| A | 6375 | hCoV-19/Luxembourg/LNS4254328/2020\|EPI_ISL_429737\| | .................... | .................... | .................... | .................... | .................... | .................... |
| A | 6376 | hCoV-19/Luxembourg/LNS5488745/2020\|EPI_ISL_429738\| | .................... | .................... | .................... | .................... | .................... | .................... |
| A | 6377 | hCoV-19/Luxembourg/LNS5549490/2020\|EPI_ISL_429740\| | .................... | .................... | .................... | .................... | .................... | .................... |
| A | 6378 | hCoV-19/Luxembourg/LNS6533667/2020\|EPI_ISL_429741\| | .................... | .................... | .................... | .................... | .................... | .................... |
| A | 6379 | hCoV-19/Luxembourg/LNS1658150/2020\|EPI_ISL_429742\| | .................... | .................... | .................... | .................... | .................... | .................... |
| A | 6380 | hCoV-19/Luxembourg/LNS6325758/2020\|EPI_ISL_429743\| | .................... | .................... | .................... | .................... | .................... | .................... |
| A | 6381 | hCoV-19/Luxembourg/LNS2913521/2020\|EPI_ISL_429746\| | .................... | .................... | .................... | .................... | .................... | .................... |
| A | 6382 | hCoV-19/Luxembourg/LNS4830703/2020\|EPI_ISL_429748\| | .................... | .................... | .................... | .................... | .................... | .................... |

| | | | | | | | | |
|---|---|---|---|---|---|---|---|---|
| A | 6383 hCoV-19/Luxembourg/LNS3989293/2020\|EPI_ISL_429749\| | ···················· | ···················· | ···················· | ···················· | ···················· | ···················· | |
| A | 6384 hCoV-19/Luxembourg/LNS9598665/2020\|EPI_ISL_429750\| | ···················· | ···················· | ···················· | ···················· | ···················· | ···················· | |
| A | 6385 hCoV-19/Luxembourg/LNS4245799/2020\|EPI_ISL_429753\| | ···················· | ···················· | ···················· | ···················· | ···················· | ···················· | |
| A | 6386 hCoV-19/Luxembourg/LNS8772889/2020\|EPI_ISL_429755\| | ···················· | ···················· | ···················· | ···················· | ···················· | ···················· | |
| A | 6387 hCoV-19/Luxembourg/LNS4946091/2020\|EPI_ISL_429756\| | ···················· | ···················· | ···················· | ···················· | ···················· | ···················· | |
| A | 6388 hCoV-19/Luxembourg/LNS7006156/2020\|EPI_ISL_429757\| | ···················· | ···················· | ···················· | ···················· | ···················· | ···················· | |
| A | 6389 hCoV-19/Luxembourg/LNS8167314/2020\|EPI_ISL_429758\| | ···················· | ···················· | ···················· | ···················· | ···················· | ···················· | |
| A | 6390 hCoV-19/Luxembourg/LNS9455951/2020\|EPI_ISL_429759\| | ···················· | ···················· | ···················· | ···················· | ···················· | ···················· | |
| A | 6391 hCoV-19/Luxembourg/LNS0211710/2020\|EPI_ISL_429760\| | ···················· | ···················· | ···················· | ···················· | ···················· | ···················· | |
| A | 6392 hCoV-19/Luxembourg/LNS6277439/2020\|EPI_ISL_429762\| | ···················· | ···················· | ···················· | ···················· | ···················· | ···················· | |
| A | 6393 hCoV-19/Luxembourg/LNS1703112/2020\|EPI_ISL_429764\| | ···················· | ···················· | ···················· | ···················· | ···················· | ···················· | |
| A | 6394 hCoV-19/Luxembourg/LNS0814393/2020\|EPI_ISL_429766\| | ···················· | ···················· | ···················· | ···················· | ···················· | ···················· | |
| A | 6395 hCoV-19/Luxembourg/LNS3437586/2020\|EPI_ISL_429767\| | ···················· | ···················· | ···················· | ···················· | ···················· | ···················· | |
| A | 6396 hCoV-19/Luxembourg/LNS7721101/2020\|EPI_ISL_429769\| | ···················· | ···················· | ···················· | ···················· | ···················· | ···················· | |
| A | 6397 hCoV-19/Luxembourg/LNS7007586/2020\|EPI_ISL_429791\| | ···················· | ···················· | ···················· | ···················· | ···················· | ···················· | |
| A | 6398 hCoV-19/Croatia/LG-S2/2020\|EPI_ISL_429792\|2020-03 | ···················· | ···················· | ···················· | ···················· | ···················· | ···················· | |
| A | 6399 hCoV-19/Luxembourg/LNS7812353/2020\|EPI_ISL_429793\| | ···················· | ···················· | ···················· | ···················· | ···················· | ···················· | |
| A | 6400 hCoV-19/Luxembourg/LNS6479831/2020\|EPI_ISL_429794\| | ···················· | ···················· | ···················· | ···················· | ···················· | ···················· | |
| A | 6401 hCoV-19/Taiwan/NTU02/2020\|EPI_ISL_410218\|2020-02-0 | ···················· | ···················· | ···················· | ···················· | ···················· | ···················· | |
| A | 6402 hCoV-19/Wuhan/HBCDC-HB-02/2019\|EPI_ISL_412898\|2019 | ···················· | ···················· | ···················· | ···················· | ···················· | ···················· | |
| A | 6403 hCoV-19/Wuhan/HBCDC-HB-03/2019\|EPI_ISL_412899\|2019 | ···················· | ···················· | ···················· | ···················· | ···················· | ···················· | |
| A | 6404 hCoV-19/Japan/DP0005/2020\|EPI_ISL_416565\|2020-02-1 | ···················· | ···················· | ···················· | ···················· | ···················· | ···················· | |
| A | 6405 hCoV-19/Japan/DP0027/2020\|EPI_ISL_416566\|2020-02-1 | ···················· | ···················· | ···················· | ···················· | ···················· | ···················· | |
| A | 6406 hCoV-19/Japan/DP0037/2020\|EPI_ISL_416567\|2020-02-1 | ···················· | ···················· | ···················· | ···················· | ···················· | ···················· | |
| A | 6407 hCoV-19/Japan/DP0059/2020\|EPI_ISL_416569\|2020-02-1 | ···················· | ···················· | ···················· | ···················· | ···················· | ···················· | |
| A | 6408 hCoV-19/Japan/DP0065/2020\|EPI_ISL_416570\|2020-02-1 | ···················· | ···················· | ···················· | ···················· | ···················· | ···················· | |
| A | 6409 hCoV-19/Japan/DP0077/2020\|EPI_ISL_416571\|2020-02-1 | ···················· | ···················· | ···················· | ···················· | ···················· | ···················· | |
| A | 6410 hCoV-19/Japan/DP0078/2020\|EPI_ISL_416572\|2020-02-1 | ···················· | ···················· | ···················· | ···················· | ···················· | ···················· | |
| A | 6411 hCoV-19/Japan/DP0104/2020\|EPI_ISL_416573\|2020-02-1 | ···················· | ···················· | ···················· | ···················· | ···················· | ···················· | |
| A | 6412 hCoV-19/Japan/DP0107/2020\|EPI_ISL_416574\|2020-02-1 | ···················· | ···················· | ···················· | ···················· | ···················· | ···················· | |
| A | 6413 hCoV-19/Japan/DP0121/2020\|EPI_ISL_416575\|2020-02-1 | ···················· | ···················· | ···················· | ···················· | ···················· | ···················· | |
| A | 6414 hCoV-19/Japan/DP0133/2020\|EPI_ISL_416576\|2020-02-1 | ···················· | ···················· | ···················· | ···················· | ···················· | ···················· | |
| A | 6415 hCoV-19/Japan/DP0134/2020\|EPI_ISL_416577\|2020-02-1 | ···················· | ···················· | ···················· | ···················· | ···················· | ···················· | |
| A | 6416 hCoV-19/Japan/DP0152/2020\|EPI_ISL_416578\|2020-02-1 | ···················· | ···················· | ···················· | ···················· | ···················· | ···················· | |
| A | 6417 hCoV-19/Japan/DP0158/2020\|EPI_ISL_416579\|2020-02-1 | ···················· | ···················· | ···················· | ···················· | ···················· | ···················· | |
| A | 6418 hCoV-19/Japan/DP0184/2020\|EPI_ISL_416580\|2020-02-1 | ···················· | ···················· | ···················· | ···················· | ···················· | ···················· | |
| A | 6419 hCoV-19/Japan/DP0190/2020\|EPI_ISL_416581\|2020-02-1 | ···················· | ···················· | ···················· | ···················· | ···················· | ···················· | |
| A | 6420 hCoV-19/Japan/DP0191/2020\|EPI_ISL_416582\|2020-02-1 | ···················· | ···················· | ···················· | ···················· | ···················· | ···················· | |
| A | 6421 hCoV-19/Japan/DP0196/2020\|EPI_ISL_416583\|2020-02-1 | ···················· | ···················· | ···················· | ···················· | ···················· | ···················· | |
| A | 6422 hCoV-19/Japan/DP0200/2020\|EPI_ISL_416584\|2020-02-1 | ···················· | ···················· | ···················· | ···················· | ···················· | ···················· | |
| A | 6423 hCoV-19/Japan/DP0236/2020\|EPI_ISL_416585\|2020-02-1 | ···················· | ···················· | ···················· | ···················· | ···················· | ···················· | |
| A | 6424 hCoV-19/Japan/DP0274/2020\|EPI_ISL_416586\|2020-02-1 | ···················· | ···················· | ···················· | ···················· | ···················· | ···················· | |
| A | 6425 hCoV-19/Japan/DP0278/2020\|EPI_ISL_416587\|2020-02-1 | ···················· | ···················· | ···················· | ···················· | ···················· | ···················· | |
| A | 6426 hCoV-19/Japan/DP0287/2020\|EPI_ISL_416589\|2020-02-1 | ···················· | ···················· | ···················· | ···················· | ···················· | ···················· | |
| A | 6427 hCoV-19/Japan/DP0289/2020\|EPI_ISL_416590\|2020-02-1 | ···················· | ···················· | ···················· | ···················· | ···················· | ···················· | |
| A | 6428 hCoV-19/Japan/DP0290/2020\|EPI_ISL_416591\|2020-02-1 | ···················· | ···················· | ···················· | ···················· | ···················· | ···················· | |
| A | 6429 hCoV-19/Japan/DP0294/2020\|EPI_ISL_416592\|2020-02-1 | ···················· | ···················· | ···················· | ···················· | ···················· | ···················· | |
| A | 6430 hCoV-19/Japan/DP0311/2020\|EPI_ISL_416593\|2020-02-1 | ···················· | ···················· | ···················· | ···················· | ···················· | ···················· | |
| A | 6431 hCoV-19/Japan/DP0319/2020\|EPI_ISL_416594\|2020-02-1 | ···················· | ···················· | ···················· | ···················· | ···················· | ···················· | |
| A | 6432 hCoV-19/Japan/DP0328/2020\|EPI_ISL_416595\|2020-02-1 | ···················· | ···················· | ···················· | ···················· | ···················· | ···················· | |
| A | 6433 hCoV-19/Japan/DP0344/2020\|EPI_ISL_416596\|2020-02-1 | ···················· | ···················· | ···················· | ···················· | ···················· | ···················· | |
| A | 6434 hCoV-19/Japan/DP0346/2020\|EPI_ISL_416597\|2020-02-1 | ···················· | ···················· | ···················· | ···················· | ···················· | ···················· | |
| A | 6435 hCoV-19/Japan/DP0357/2020\|EPI_ISL_416598\|2020-02-1 | ···················· | ···················· | ···················· | ···················· | ···················· | ···················· | |
| A | 6436 hCoV-19/Japan/DP0361/2020\|EPI_ISL_416599\|2020-02-1 | ···················· | ···················· | ···················· | ···················· | ···················· | ···················· | |
| A | 6437 hCoV-19/Wales/PHWC-26A7F/2020\|EPI_ISL_422200\|2020- | ···················· | ···A················ | ···················· | ···················· | ···················· | ···················· | |
| A | 6438 hCoV-19/Wales/PHWC-25EA1/2020\|EPI_ISL_422202\|2020- | ···················· | ···················· | ···················· | ···················· | ···················· | ···················· | |
| A | 6439 hCoV-19/Wales/PHWC-26F16/2020\|EPI_ISL_422204\|2020- | ···················· | ···················· | ···················· | ···················· | ···················· | ···················· | |
| A | 6440 hCoV-19/Wales/PHWC-268DF/2020\|EPI_ISL_422209\|2020- | ···················· | ···················· | ···················· | ···················· | ···················· | ···················· | |
| A | 6441 hCoV-19/Wales/PHWC-26F61/2020\|EPI_ISL_422214\|2020- | ···················· | ···················· | ···················· | ···················· | ···················· | ···················· | |
| A | 6442 hCoV-19/Wales/PHWC-26E0A/2020\|EPI_ISL_422215\|2020- | ···················· | ···M················ | ···················· | ···················· | ···················· | ···················· | |
| A | 6443 hCoV-19/Taiwan/NTU04/2020\|EPI_ISL_422407\|2020-03-0 | ···················· | ···················· | ···················· | ···················· | ···················· | ···················· | |
| A | 6444 hCoV-19/Taiwan/NTU05/2020\|EPI_ISL_422408\|2020-03-1 | ···················· | ···················· | ···················· | ···················· | ···················· | ···················· | |
| A | 6445 hCoV-19/Taiwan/NTU06/2020\|EPI_ISL_422409\|2020-03-1 | ···················· | ···················· | ···················· | ···················· | ···················· | ···················· | |
| A | 6446 hCoV-19/Taiwan/NTU07/2020\|EPI_ISL_422410\|2020-03-1 | ···················· | ···················· | ···················· | ···················· | ···················· | ···················· | |
| A | 6447 hCoV-19/Taiwan/NTU08/2020\|EPI_ISL_422411\|2020-03-1 | ···················· | ···················· | ···················· | ···················· | ···················· | ···················· | |
| A | 6448 hCoV-19/Taiwan/NTU09/2020\|EPI_ISL_422412\|2020-03-1 | ···················· | ···················· | ···················· | ···················· | ···················· | ···················· | |
| A | 6449 hCoV-19/Taiwan/NTU11/2020\|EPI_ISL_422413\|2020-03-1 | ···················· | ···················· | ···················· | ···················· | ···················· | ···················· | |
| A | 6450 hCoV-19/Taiwan/NTU12/2020\|EPI_ISL_422414\|2020-03-1 | ···················· | ···················· | ···················· | ···················· | ···················· | ···················· | |
| A | 6451 hCoV-19/Taiwan/NTU13/2020\|EPI_ISL_422415\|2020-03-1 | ···················· | ···················· | ···················· | ···················· | ···················· | ···················· | |
| A | 6452 hCoV-19/Taiwan/NTU14/2020\|EPI_ISL_422416\|2020-03-1 | ···················· | ···················· | ···················· | ···················· | ···················· | ···················· | |
| A | 6453 hCoV-19/Taiwan/NTU15/2020\|EPI_ISL_422417\|2020-03-1 | ···················· | ···················A | ···················· | ···················· | ···················· | ···················· | 1 |
| A | 6454 hCoV-19/Taiwan/NTU16/2020\|EPI_ISL_422418\|2020-03-1 | ···················· | ···················· | ···················· | ···················· | ···················· | ···················· | |
| A | 6455 hCoV-19/Taiwan/NTU17/2020\|EPI_ISL_422419\|2020-03-1 | ···················· | ···················· | ···················· | ···················· | ···················· | ···················· | |
| A | 6456 hCoV-19/Taiwan/NTU19/2020\|EPI_ISL_422420\|2020-03-1 | ···················· | ···················· | ···················· | ···················· | ···················· | ···················· | |
| A | 6457 hCoV-19/Taiwan/NTU20/2020\|EPI_ISL_422421\|2020-03-1 | ···················· | ···················· | ···················· | ···················· | ···················· | ···················· | |
| A | 6458 hCoV-19/Taiwan/NTU21/2020\|EPI_ISL_422422\|2020-03-1 | ···················· | ···················· | ···················· | ···················· | ···················· | ···················· | |
| A | 6459 hCoV-19/Kuwait/KU006/2020\|EPI_ISL_422424\|2020-03-1 | ···················· | ···················· | ···················· | ···················· | ···················· | ···················· | |
| A | 6460 hCoV-19/Zhejiang/HZ103/2020\|EPI_ISL_422425\|2020-01 | ···················· | ···················· | ···················· | ···················· | ···················· | ···················· | |

| | | | | | | | |
|---|---|---|---|---|---|---|---|
| A | 6461 hCoV-19/Kuwait/KU005/2020\|EPI_ISL_422426\|2020-03-1 | ···················· | ···················· | ···················· | ···················· | ···················· | ···················· |
| A | 6462 hCoV-19/Kuwait/KU008/2020\|EPI_ISL_422427\|2020-03-1 | ···················· | ···················· | ···················· | ···················· | ···················· | ···················· |
| A | 6463 hCoV-19/Singapore/35/2020\|EPI_ISL_422428\|2020-02-0 | ···················· | ···················· | ···················· | ···················· | ···················· | ···················· |
| A | 6464 hCoV-19/Singapore/36/2020\|EPI_ISL_422429\|2020-02-1 | ···················· | ···················· | ···················· | ···················· | ···················· | ···················· |
| A | 6465 hCoV-19/Singapore/37/2020\|EPI_ISL_422430\|2020-03-1 | ···················· | ···················· | ···················· | ···················· | ···················· | ···················· |
| A | 6466 hCoV-19/Singapore/38/2020\|EPI_ISL_422431\|2020-03-3 | ···················· | ···················· | ···················· | ···················· | ···················· | ···················· |
| A | 6467 hCoV-19/Singapore/39/2020\|EPI_ISL_422432\|2020-03-3 | ···················· | ···················· | ···················· | ···················· | ···················· | ···················· |
| A | 6468 hCoV-19/Singapore/40/2020\|EPI_ISL_422433\|2020-03-3 | ···················· | ···················· | ···················· | ···················· | ···················· | ···················· |
| A | 6469 hCoV-19/Singapore/41/2020\|EPI_ISL_422434\|2020-03-3 | ···················· | ···················· | ···················· | ···················· | ···················· | ···················· |
| A | 6472 hCoV-19/England/20144024304/2020\|EPI_ISL_423532\|20 | ···················· | ···················· | ···················· | ···················· | ···················· | ···················· |
| A | 6473 hCoV-19/England/20144036004/2020\|EPI_ISL_423535\|20 | ···················· | ···················· | ···················· | ···················· | ···················· | ···················· |
| A | 6475 hCoV-19/England/20144061104/2020\|EPI_ISL_423550\|20 | ···················· | ···················· | ···················· | ···················· | ···················· | ···················· |
| A | 6476 hCoV-19/England/20144064004/2020\|EPI_ISL_423554\|20 | ···················· | ···················· | ···················· | ···················· | ···················· | ···················· |
| A | 6477 hCoV-19/England/20144066304/2020\|EPI_ISL_423555\|20 | ···T················ | ···················· | ···················· | ···················· | ···················· | ···················· |
| A | 6479 hCoV-19/England/20144071504/2020\|EPI_ISL_423559\|20 | ···················· | ···················· | ···················· | ···················· | ···················· | ···················· |
| A | 6480 hCoV-19/England/20144073804/2020\|EPI_ISL_423561\|20 | ···················· | ···················· | ···················· | ···················· | ···················· | ···················· |
| A | 6481 hCoV-19/England/20144074504/2020\|EPI_ISL_423563\|20 | ···················· | ···················· | ···················· | ···················· | ···················· | ···················· |
| A | 6483 hCoV-19/England/20144076204/2020\|EPI_ISL_423570\|20 | ···················· | ···················· | ···················· | ···················· | ···················· | ···················· |
| A | 6484 hCoV-19/England/20146004804/2020\|EPI_ISL_423573\|20 | ···················· | ···················· | ···················· | ···················· | ···················· | ···················· |
| A | 6485 hCoV-19/England/20146005804/2020\|EPI_ISL_423574\|20 | ···················· | ···················· | ···················· | ···················· | ···················· | ···················· |
| A | 6488 hCoV-19/England/20146025304/2020\|EPI_ISL_423582\|20 | ···················· | ···················· | ···················· | ···················· | ···················· | ···················· |
| A | 6489 hCoV-19/England/20146060504/2020\|EPI_ISL_423583\|20 | ···················· | ···················· | ···················· | ···················· | ···················· | ···················· |
| A | 6492 hCoV-19/England/20148006604/2020\|EPI_ISL_423588\|20 | ···················· | ···················· | ···················· | ···················· | ···················· | ···················· |
| A | 6494 hCoV-19/England/20148043704/2020\|EPI_ISL_423591\|20 | ···················· | ···················· | ···················· | ···················· | ···················· | ···················· |
| A | 6496 hCoV-19/England/20132001104/2020\|EPI_ISL_423704\|20 | ···················· | ···················· | ···················· | ···················· | ···················· | ···················· |
| A | 6497 hCoV-19/England/20132003004/2020\|EPI_ISL_423711\|20 | ···················· | ···················· | ···················· | ···················· | ···················· | ···················· |
| A | 6498 hCoV-19/England/20132003204/2020\|EPI_ISL_423712\|20 | ···················· | ···················· | ···················· | ···················· | ···················· | ···················· |
| A | 6500 hCoV-19/England/20132004804/2020\|EPI_ISL_423717\|20 | ···················· | ···················· | ···················· | ···················· | ··N········N········ | ···················· |
| A | 6501 hCoV-19/England/20132004904/2020\|EPI_ISL_423718\|20 | ···················· | ···················· | ···················· | ···················· | ···················· | ···················· |
| A | 6502 hCoV-19/England/20132005204/2020\|EPI_ISL_423720\|20 | ···················· | ···················· | ···················· | ···················· | ···················· | ···················· |
| A | 6503 hCoV-19/England/20132005704/2020\|EPI_ISL_423723\|20 | ···················· | ···················· | ···················· | ···················· | ···················· | ···················· |
| A | 6504 hCoV-19/England/20132017604/2020\|EPI_ISL_423727\|20 | ···················· | ···················· | ···················· | ···················· | ···················· | ···················· |
| A | 6505 hCoV-19/England/20132018004/2020\|EPI_ISL_423731\|20 | ···················· | ···················· | ···················· | ···················· | ···················· | ···················· |
| A | 6507 hCoV-19/England/20132018704/2020\|EPI_ISL_423733\|20 | ···················· | ···················· | ···················· | ···················· | ···················· | ···················· |
| A | 6508 hCoV-19/England/20132018804/2020\|EPI_ISL_423734\|20 | ···················· | ···················· | ···················· | ···················· | ···················· | ···················· |
| A | 6509 hCoV-19/England/20132018904/2020\|EPI_ISL_423735\|20 | ···················· | ···················· | ···················· | ···················· | ···················· | ···················· |
| A | 6511 hCoV-19/England/20132021004/2020\|EPI_ISL_423741\|20 | ···················· | ···················· | ···················· | ···················· | ···················· | ···················· |
| A | 6512 hCoV-19/England/20132023204/2020\|EPI_ISL_423749\|20 | ···················· | ···················· | ···················· | ···················· | ···················· | ···················· |
| A | 6513 hCoV-19/England/20132023704/2020\|EPI_ISL_423752\|20 | ···················· | ···················· | ···················· | ···················· | ···················· | ···················· |
| A | 6514 hCoV-19/England/20132023904/2020\|EPI_ISL_423753\|20 | ···················· | ···················· | ···················· | ···················· | ···················· | ···················· |
| A | 6515 hCoV-19/England/20132024604/2020\|EPI_ISL_423758\|20 | ···················· | ···················· | ···················· | ···················· | ···················· | ···················· |
| A | 6516 hCoV-19/England/20132026904/2020\|EPI_ISL_423761\|20 | ···················· | ···················· | ···················· | ···················· | ···················· | ···················· |
| A | 6517 hCoV-19/England/20132027004/2020\|EPI_ISL_423762\|20 | ···················· | ···················· | ···················· | ···················· | ···················· | ···················· |
| A | 6518 hCoV-19/England/20132027504/2020\|EPI_ISL_423765\|20 | ···················· | ···················· | ···················· | ···················· | ···················· | ···················· |
| A | 6519 hCoV-19/USA/FL_7169/2020\|EPI_ISL_424855\|2020-03-02 | ···················· | ···················· | ···················· | ···················· | ···················· | ···················· |
| A | 6520 hCoV-19/USA/FL_9656/2020\|EPI_ISL_424856\|2020-03-06 | ···················· | ···················· | ···················· | ···················· | ···················· | ···················· |
| A | 6521 hCoV-19/USA/FL_1177/2020\|EPI_ISL_424857\|2020-03-06 | ···················· | ···················· | ···················· | ···················· | ···················· | ···················· |
| A | 6522 hCoV-19/USA/GA_1621/2020\|EPI_ISL_424858\|2020-03-04 | ···················· | ···················· | ···················· | ···················· | ···················· | ···················· |
| A | 6523 hCoV-19/USA/GA_1847/2020\|EPI_ISL_424859\|2020-03-05 | ···················· | ···················· | ···················· | ···················· | ···················· | ···················· |
| A | 6524 hCoV-19/USA/GA_2055/2020\|EPI_ISL_424860\|2020-03 | ···················· | ···················· | ···················· | ···················· | ···················· | ···················· |
| A | 6525 hCoV-19/USA/GA_2118/2020\|EPI_ISL_424861\|2020-03-08 | ···················· | ···················· | ···················· | ···················· | ···················· | ···················· |
| A | 6526 hCoV-19/USA/GA_2120/2020\|EPI_ISL_424862\|2020-03 | ···················· | ···················· | ···················· | ···················· | ···················· | ···················· |
| A | 6527 hCoV-19/USA/GA_2185/2020\|EPI_ISL_424863\|2020-03 | ···················· | ···················· | ···················· | ···················· | ···················· | ···················· |
| A | 6528 hCoV-19/USA/GA_2343/2020\|EPI_ISL_424864\|2020-03-10 | ···················· | ···················· | ···················· | ···················· | ···················· | ···················· |
| A | 6529 hCoV-19/USA/IA_6394/2020\|EPI_ISL_424865\|2020-03-08 | ···················· | ···················· | ···················· | ···················· | ···················· | ···················· |
| A | 6530 hCoV-19/USA/IN_2001/2020\|EPI_ISL_424866\|2020-03-09 | ···················· | ···················· | ···················· | ···················· | ···················· | ···················· |
| A | 6531 hCoV-19/USA/KS_4126/2020\|EPI_ISL_424867\|2020-03-06 | ···················· | ···················· | ···················· | ···················· | ···················· | ···················· |
| A | 6532 hCoV-19/USA/LA_0842/2020\|EPI_ISL_424868\|2020-03-09 | ···················· | ···················· | ···················· | ···················· | ···················· | ···················· |
| A | 6533 hCoV-19/USA/MD_0026/2020\|EPI_ISL_424869\|2020-03-04 | ···················· | ···················· | ···················· | ···················· | ···················· | ···················· |
| A | 6534 hCoV-19/USA/MO_4470/2020\|EPI_ISL_424870\|2020-03-06 | ···················· | ···················· | ···················· | ···················· | ···················· | ···················· |
| A | 6535 hCoV-19/USA/NC_6999/2020\|EPI_ISL_424871\|2020-03-06 | ···················· | ···················· | ···················· | ···················· | ···················· | ···················· |
| A | 6536 hCoV-19/USA/NC_0024/2020\|EPI_ISL_424872\|2020-03-08 | ···················· | ···················· | ···················· | ···················· | ···················· | ···················· |
| A | 6537 hCoV-19/USA/NC_0031/2020\|EPI_ISL_424873\|2020-03-07 | ···················· | ···················· | ···················· | ···················· | ···················· | ···················· |
| A | 6538 hCoV-19/USA/NE_6605/2020\|EPI_ISL_424874\|2020-03-05 | ···················· | ···················· | ···················· | ···················· | ···················· | ···················· |
| A | 6539 hCoV-19/USA/NE_7025/2020\|EPI_ISL_424875\|2020-03-07 | ···················· | ···················· | ···················· | ···················· | ···················· | ···················· |
| A | 6540 hCoV-19/USA/NH_0029/2020\|EPI_ISL_424876\|2020-03-06 | ···················· | ···················· | ···················· | ···················· | ···················· | ···················· |
| A | 6541 hCoV-19/USA/NH_0033/2020\|EPI_ISL_424877\|2020-03-06 | ···················· | ···················· | ···················· | ···················· | ···················· | ···················· |
| A | 6542 hCoV-19/USA/NJ_3592/2020\|EPI_ISL_424878\|2020-03-04 | ···················· | ···················· | ···················· | ···················· | ···················· | ···················· |
| A | 6543 hCoV-19/USA/NV_0052/2020\|EPI_ISL_424879\|2020-03-09 | ···················· | ···················· | ···················· | ···················· | ···················· | ···················· |
| A | 6544 hCoV-19/USA/OH_0023/2020\|EPI_ISL_424880\|2020-03-08 | ···················· | ···················· | ···················· | ···················· | ···················· | ···················· |
| A | 6545 hCoV-19/USA/PA_1802/2020\|EPI_ISL_424881\|2020-03-05 | ···················· | ···················· | ···················· | ···················· | ···················· | ···················· |
| A | 6546 hCoV-19/USA/PA_1881/2020\|EPI_ISL_424882\|2020-03-05 | ···················· | ···················· | ···················· | ···················· | ···················· | ···················· |
| A | 6547 hCoV-19/USA/PA_2317/2020\|EPI_ISL_424883\|2020-03-07 | ···················· | ···················· | ···················· | ···················· | ···················· | ···················· |
| A | 6548 hCoV-19/USA/PA_2937/2020\|EPI_ISL_424884\|2020-03-08 | ···················· | ···················· | ···················· | ···················· | ···················· | ···················· |
| A | 6549 hCoV-19/USA/PA_4395/2020\|EPI_ISL_424885\|2020-03-09 | ···················· | ···················· | ···················· | ···················· | ···················· | ···················· |
| A | 6550 hCoV-19/USA/PA_4405/2020\|EPI_ISL_424886\|2020-03-06 | ···················· | ···················· | ···················· | ···················· | ···················· | ···················· |
| A | 6551 hCoV-19/USA/RI_0702/2020\|EPI_ISL_424887\|2020-03-05 | ···················· | ···················· | ···················· | ···················· | ···················· | ···················· |
| A | 6552 hCoV-19/USA/SC_6370/2020\|EPI_ISL_424888\|2020-03-08 | ···················· | ···················· | ···················· | ···················· | ···················· | ···················· |

| | | | | | | | | |
|---|---|---|---|---|---|---|---|---|
| A | 6553 | hCoV-19/USA/UT_8199/2020\|EPI_ISL_424889\|2020-03-05 | .................... | .................... | .................... | .................... | .................... | .................... |
| A | 6554 | hCoV-19/USA/UT_8906/2020\|EPI_ISL_424890\|2020-03-07 | .................... | .................... | .................... | .................... | .................... | .................... |
| A | 6555 | hCoV-19/USA/VA_6352/2020\|EPI_ISL_424891\|2020-03-08 | .................... | .................... | .................... | .................... | .................... | .................... |
| A | 6556 | hCoV-19/USA/VA_6377/2020\|EPI_ISL_424892\|2020-03-08 | .................... | .................... | .................... | .................... | .................... | .................... |
| A | 6557 | hCoV-19/USA/VA_6382/2020\|EPI_ISL_424893\|2020-03-08 | .................... | .................... | .................... | .................... | .................... | .................... |
| A | 6558 | hCoV-19/USA/VA_6389/2020\|EPI_ISL_424894\|2020-03-08 | .................... | .................... | .................... | .................... | .................... | .................... |
| A | 6559 | hCoV-19/USA/IA_6396/2020\|EPI_ISL_424895\|2020-03-08 | .................... | .................... | .................... | .................... | .................... | .................... |
| A | 6560 | hCoV-19/USA/IA_6395/2020\|EPI_ISL_424896\|2020-03-08 | .................... | .................... | .................... | .................... | .................... | .................... |
| A | 6561 | hCoV-19/USA/IA_6401/2020\|EPI_ISL_424897\|2020-03-08 | .................... | .................... | .................... | .................... | .................... | .................... |
| A | 6562 | hCoV-19/USA/IA_6399/2020\|EPI_ISL_424898\|2020-03-08 | .................... | .................... | .................... | .................... | .................... | .................... |
| A | 6563 | hCoV-19/USA/IA_6391/2020\|EPI_ISL_424899\|2020-03-07 | .................... | .................... | .................... | .................... | .................... | .................... |
| A | 6564 | hCoV-19/USA/WA-UW-3918/2020\|EPI_ISL_427209\|2020-03 | .................... | .................... | .................... | .................... | .................... | .................... |
| A | 6565 | hCoV-19/USA/WA-UW-3923/2020\|EPI_ISL_427210\|2020-03 | .................... | .................... | .................... | .................... | .................... | .................... |
| A | 6566 | hCoV-19/USA/WA-UW-3932/2020\|EPI_ISL_427211\|2020-03 | .................... | .................... | .................... | .................... | .................... | .................... |
| A | 6567 | hCoV-19/USA/WA-UW-3935/2020\|EPI_ISL_427212\|2020-03 | .................... | .................... | .................... | .................... | .................... | .................... |
| A | 6568 | hCoV-19/USA/WA-UW-3939/2020\|EPI_ISL_427213\|2020-03 | .................... | .................... | .................... | .................... | .................... | .................... |
| A | 6569 | hCoV-19/USA/WA-UW-3940/2020\|EPI_ISL_427214\|2020-03 | .................... | .................... | .................... | .................... | .................... | .................... |
| A | 6570 | hCoV-19/USA/WA-UW-3941/2020\|EPI_ISL_427215\|2020-03 | .................... | .................... | .................... | .................... | .................... | .................... |
| A | 6571 | hCoV-19/USA/WA-UW-3942/2020\|EPI_ISL_427216\|2020-03 | .................... | .................... | .................... | .................... | .................... | .................... |
| A | 6572 | hCoV-19/USA/WA-UW-3946/2020\|EPI_ISL_427217\|2020-03 | .................... | .................... | .................... | .................... | .................... | .................... |
| A | 6573 | hCoV-19/USA/WA-UW-3950/2020\|EPI_ISL_427218\|2020-03 | .................... | .................... | .................... | .................... | .................... | .................... |
| A | 6574 | hCoV-19/USA/WA-UW-3954/2020\|EPI_ISL_427219\|2020-03 | .................... | .................... | .................... | .................... | .................... | .................... |
| A | 6575 | hCoV-19/USA/WA-UW-3959/2020\|EPI_ISL_427220\|2020-03 | .................... | .................... | .................... | .................... | .................... | .................... |
| A | 6576 | hCoV-19/USA/WA-UW-3964/2020\|EPI_ISL_427222\|2020-03 | .................... | .................... | .................... | .................... | .................... | .................... |
| A | 6577 | hCoV-19/USA/WA-UW-3969/2020\|EPI_ISL_427223\|2020-03 | .................... | .................... | .................... | .................... | .................... | .................... |
| A | 6578 | hCoV-19/USA/WA-UW-3983/2020\|EPI_ISL_427224\|2020-03 | .................... | .................... | .................... | .................... | .................... | .................... |
| A | 6579 | hCoV-19/USA/WA-UW-3984/2020\|EPI_ISL_427225\|2020-03 | .................... | .................... | .................... | .................... | .................... | .................... |
| A | 6580 | hCoV-19/USA/WA-UW-3985/2020\|EPI_ISL_427226\|2020-03 | .................... | .................... | .................... | .................... | .................... | .................... |
| A | 6581 | hCoV-19/USA/WA-UW-3986/2020\|EPI_ISL_427227\|2020-03 | .................... | .................... | .................... | .................... | .................... | .................... |
| A | 6582 | hCoV-19/USA/WA-UW-3988/2020\|EPI_ISL_427228\|2020-03 | .................... | .................... | .................... | .................... | .................... | .................... |
| A | 6583 | hCoV-19/USA/WA-UW-3989/2020\|EPI_ISL_427229\|2020-03 | .................... | .................... | .................... | .................... | .................... | .................... |
| A | 6584 | hCoV-19/USA/WA-UW-3990/2020\|EPI_ISL_427230\|2020-03 | .................... | .................... | .................... | .................... | .................... | .................... |
| A | 6585 | hCoV-19/USA/WA-UW-3994/2020\|EPI_ISL_427232\|2020-03 | .................... | .................... | .................... | .................... | .................... | .................... |
| A | 6586 | hCoV-19/USA/WA-UW-3996/2020\|EPI_ISL_427233\|2020-03 | .................... | .................... | .................... | .................... | .................... | .................... |
| A | 6587 | hCoV-19/USA/WA-UW-3997/2020\|EPI_ISL_427234\|2020-03 | .................... | .................... | .................... | .................... | .................... | .................... |
| A | 6588 | hCoV-19/USA/WA-UW-4035/2020\|EPI_ISL_427235\|2020-03 | .................... | .................... | .................... | .................... | .................... | .................... |
| A | 6589 | hCoV-19/USA/WA-UW-4038/2020\|EPI_ISL_427236\|2020-03 | .................... | .................... | .................... | .................... | .................... | .................... |
| A | 6590 | hCoV-19/USA/WA-UW-4039/2020\|EPI_ISL_427237\|2020-03 | .................... | .................... | .................... | .................... | .................... | .................... |
| A | 6591 | hCoV-19/USA/WA-UW-4068/2020\|EPI_ISL_427238\|2020-04 | .................... | .................... | .................... | .................... | .................... | .................... |
| A | 6592 | hCoV-19/USA/WA-UW-4082/2020\|EPI_ISL_427239\|2020-04 | .................... | .................... | .................... | .................... | .................... | .................... |
| A | 6593 | hCoV-19/USA/WA-UW-4105/2020\|EPI_ISL_427240\|2020-03 | .................... | .................... | .................... | .................... | .................... | .................... |
| A | 6594 | hCoV-19/USA/WA-UW-4109/2020\|EPI_ISL_427241\|2020-03 | .................... | .................... | .................... | .................... | .................... | .................... |
| A | 6595 | hCoV-19/USA/WA-UW-4111/2020\|EPI_ISL_427242\|2020-03 | .................... | .................... | .................... | .................... | .................... | .................... |
| A | 6596 | hCoV-19/USA/WA-UW-4180/2020\|EPI_ISL_427245\|2020-04 | .................... | .................... | .................... | .................... | .................... | .................... |
| A | 6597 | hCoV-19/USA/WA-UW-4188/2020\|EPI_ISL_427246\|2020-03 | .................... | .................... | .................... | .................... | .................... | .................... |
| A | 6598 | hCoV-19/USA/WA-UW-4197/2020\|EPI_ISL_427247\|2020-03 | .................... | .................... | .................... | .................... | .................... | .................... |
| A | 6599 | hCoV-19/USA/WA-UW-4213/2020\|EPI_ISL_427248\|2020-03 | .................... | .................... | .................... | .................... | .................... | .................... |
| A | 6600 | hCoV-19/USA/WA-UW-4217/2020\|EPI_ISL_427249\|2020-03 | .................... | .................... | .................... | .................... | .................... | .................... |
| A | 6601 | hCoV-19/USA/WA-UW-4227/2020\|EPI_ISL_427250\|2020-03 | .................... | .................... | .................... | .................... | .................... | .................... |
| A | 6602 | hCoV-19/USA/WA-UW-4228/2020\|EPI_ISL_427251\|2020-03 | .................... | .................... | .................... | .................... | .................... | .................... |
| A | 6603 | hCoV-19/USA/WA-UW-4281/2020\|EPI_ISL_427252\|2020-03 | .................... | .................... | .................... | .................... | .................... | .................... |
| A | 6604 | hCoV-19/USA/WA-UW-4289/2020\|EPI_ISL_427253\|2020-03 | .................... | .................... | .................... | .................... | .................... | .................... |
| A | 6605 | hCoV-19/USA/WA-UW-4290/2020\|EPI_ISL_427254\|2020-03 | .................... | .................... | .................... | .................... | .................... | .................... |
| A | 6606 | hCoV-19/USA/WA-UW-4500/2020\|EPI_ISL_427270\|2020-03 | .................... | .................... | .................... | .................... | .................... | .................... |
| A | 6607 | hCoV-19/USA/AZ-TG271866/2020\|EPI_ISL_427271\|2020-0 | .................... | .................... | .................... | .................... | .................... | .................... |
| A | 6608 | hCoV-19/USA/AZ-TG271868/2020\|EPI_ISL_427272\|2020-0 | .................... | .................... | .................... | .................... | .................... | .................... |
| A | 6609 | hCoV-19/USA/MN-MDH-117/2020\|EPI_ISL_427273\|2020-03 | .................... | .................... | .................... | .................... | .................... | .................... |
| A | 6610 | hCoV-19/USA/MN-MDH-118/2020\|EPI_ISL_427274\|2020-03 | .................... | .................... | .................... | .................... | .................... | .................... |
| A | 6611 | hCoV-19/USA/MN-MDH-119/2020\|EPI_ISL_427275\|2020-03 | .................... | .................... | .................... | .................... | .................... | .................... |
| A | 6612 | hCoV-19/USA/MN-MDH-120/2020\|EPI_ISL_427276\|2020-03 | .................... | .................... | .................... | .................... | .................... | .................... |
| A | 6613 | hCoV-19/USA/MN-MDH-121/2020\|EPI_ISL_427277\|2020-03 | .................... | .................... | .................... | .................... | .................... | .................... |
| A | 6614 | hCoV-19/USA/MN-MDH-138/2020\|EPI_ISL_427278\|2020-03 | .................... | .................... | .................... | .................... | .................... | .................... |
| A | 6615 | hCoV-19/USA/MN-MDH-140/2020\|EPI_ISL_427279\|2020-03 | .................... | .................... | .................... | .................... | .................... | .................... |
| A | 6616 | hCoV-19/USA/MN-MDH-141/2020\|EPI_ISL_427280\|2020-03 | .................... | .................... | .................... | .................... | .................... | .................... |
| A | 6617 | hCoV-19/USA/MN-MDH-142/2020\|EPI_ISL_427281\|2020-03 | .................... | .................... | .................... | .................... | .................... | .................... |
| A | 6618 | hCoV-19/USA/MN-MDH-143/2020\|EPI_ISL_427282\|2020-03 | .................... | .................... | .................... | .................... | .................... | .................... |
| A | 6619 | hCoV-19/USA/MN-MDH-144/2020\|EPI_ISL_427283\|2020-03 | .................... | .................... | .................... | .................... | .................... | .................... |
| A | 6620 | hCoV-19/USA/MN-MDH-145/2020\|EPI_ISL_427284\|2020-03 | .................... | .................... | .................... | .................... | .................... | .................... |
| A | 6621 | hCoV-19/USA/MN-MDH-146/2020\|EPI_ISL_427285\|2020-03 | .................... | .................... | .................... | .................... | .................... | .................... |
| A | 6622 | hCoV-19/USA/MN-MDH-147/2020\|EPI_ISL_427286\|2020-03 | .................... | .................... | .................... | .................... | .................... | .................... |
| A | 6623 | hCoV-19/USA/MN-MDH-149/2020\|EPI_ISL_427287\|2020-03 | .................... | .................... | .................... | .................... | .................... | .................... |
| A | 6624 | hCoV-19/USA/OH-OSUP0016/2020\|EPI_ISL_427288\|2020-0 | .................... | .................... | .................... | .................... | .................... | .................... |
| A | 6625 | hCoV-19/USA/OH-OSUP0017/2020\|EPI_ISL_427290\|2020-0 | .................... | .................... | .................... | .................... | .................... | .................... |
| A | 6626 | hCoV-19/Brazil/AL-837/2020\|EPI_ISL_427292\|2020-03- | .................... | .................... | .................... | .................... | .................... | .................... |
| A | 6627 | hCoV-19/Brazil/BA-510/2020\|EPI_ISL_427293\|2020-03- | .................... | .................... | .................... | .................... | .................... | .................... |
| A | 6628 | hCoV-19/Brazil/DF-615i/2020\|EPI_ISL_427294\|2020-03 | .................... | .................... | .................... | .................... | .................... | .................... |
| A | 6629 | hCoV-19/Brazil/DF-619i/2020\|EPI_ISL_427295\|2020-03 | .................... | .................... | .................... | .................... | .................... | .................... |
| A | 6630 | hCoV-19/Brazil/DF-861/2020\|EPI_ISL_427296\|2020-03- | .................... | .................... | .................... | .................... | .................... | .................... |

| | | | | | | | | | |
|---|---|---|---|---|---|---|---|---|---|
| A | 6631 | hCoV-19/Brazil/DF-862/2020\|EPI_ISL_427297\|2020-03- | ···················· | ···················· | ···················· | ···················· | ···················· | ···················· | |
| A | 6632 | hCoV-19/Brazil/DF-891/2020\|EPI_ISL_427298\|2020-03- | ···················· | ···················· | ···················· | ···················· | ···················· | ···················· | |
| A | 6633 | hCoV-19/Brazil/RJ-352/2020\|EPI_ISL_427299\|2020-03- | ···················· | ···················· | ···················· | ···················· | ···················· | ···················· | |
| A | 6634 | hCoV-19/Luxembourg/LNS6137379/2020\|EPI_ISL_429722\| | ···················· | ···················· | ···················· | ···················· | ···················· | ···················· | |
| A | 6635 | hCoV-19/Luxembourg/LNS9984061/2020\|EPI_ISL_429724\| | ···················· | ···················· | ···················· | ···················· | ···················· | ···················· | |
| A | 6636 | hCoV-19/Luxembourg/LNS0756265/2020\|EPI_ISL_429725\| | ···················· | ·A·················· | ···················· | ···················· | ···················· | ···················· | |
| A | 6637 | hCoV-19/Luxembourg/LNS9102295/2020\|EPI_ISL_429727\| | ···················· | ···················· | ···················· | ···················· | ···················· | ···················· | |
| A | 6638 | hCoV-19/Turkey/HSGM-8990/2020\|EPI_ISL_429870\|2020- | ···················· | ···················· | ···················· | ···················· | ···················· | ···················· | |
| A | 6639 | hCoV-19/Turkey/HSGM-10241/2020\|EPI_ISL_429871\|2020 | ···················· | ···················· | ···················· | ···················· | ···················· | ···················· | |
| A | 6640 | hCoV-19/Turkey/HSGM-11518/2020\|EPI_ISL_429872\|2020 | ···················· | ···················· | ···················· | ···················· | ···················· | ···················· | |
| A | 6641 | hCoV-19/Turkey/HSGM-11520/2020\|EPI_ISL_429873\|2020 | ···················· | ···················· | ···················· | ···················· | ···················· | ···················· | |
| A | 6642 | hCoV-19/USA/CA-SCCPHD-UC34/2020\|EPI_ISL_429879\|202 | ···················· | ···················· | ···················· | ···················· | ···················· | NNN················· | 1 |
| A | 6643 | hCoV-19/Taiwan/299/2020\|EPI_ISL_429882\|2020-03-29 | ···················· | ···················· | ···················· | ···················· | ···················· | ···················· | |
| A | 6644 | hCoV-19/Taiwan/108/2020\|EPI_ISL_429883\|2020-03-23 | ···················· | ···················· | ···················· | ···················· | ···················· | ···················· | |
| A | 6645 | hCoV-19/Taiwan/284/2020\|EPI_ISL_429884\|2020-03-30 | ···················· | ···················· | ···················· | ···················· | ···················· | ···················· | |
| A | 6646 | hCoV-19/USA/WA-UW398/2020\|EPI_ISL_529200\|2020-03-2 | ···················· | ···················· | ···················· | ···················· | ···················· | ···················· | |
| A | 6647 | hCoV-19/USA/WA-UW391/2020\|EPI_ISL_529208\|2020-03-3 | ···················· | ···················· | ···················· | ···················· | ···················· | ···················· | |
| A | 6648 | hCoV-19/USA/WA-UW392/2020\|EPI_ISL_529209\|2020-03-2 | ···················· | ···················· | ···················· | ···················· | ···················· | ···················· | |
| A | 6649 | hCoV-19/USA/WA-UW408/2020\|EPI_ISL_529210\|2020-03-3 | ···················· | ···················· | ···················· | ···················· | ···················· | ···················· | |
| A | 6650 | hCoV-19/USA/WA-UW410/2020\|EPI_ISL_529211\|2020-03-3 | ···················· | ···················· | ···················· | ···················· | ···················· | ···················· | |
| A | 6651 | hCoV-19/USA/WA-UW413/2020\|EPI_ISL_529212\|2020-03-2 | ···················· | ···················· | ···················· | ···················· | ···················· | ···················· | |
| A | 6652 | hCoV-19/Wuhan/IME-WH01/2020\|EPI_ISL_529213\|2019-12 | ···················· | ···················· | ···················· | ···················· | ···················· | ···················· | |
| A | 6653 | hCoV-19/Wuhan/IME-WH02/2020\|EPI_ISL_529214\|2019-12 | ···················· | ···················· | ···················· | ···················· | ···················· | ···················· | |
| A | 6654 | hCoV-19/Wuhan/IME-WH03/2020\|EPI_ISL_529215\|2019-12 | ···················· | ···················· | ···················· | ···················· | ···················· | ···················· | |
| A | 6655 | hCoV-19/Wuhan/IME-WH04/2020\|EPI_ISL_529216\|2019-12 | ···················· | ···················· | ···················· | ···················· | ···················· | ···················· | |
| A | 6656 | hCoV-19/Wuhan/IME-WH05/2020\|EPI_ISL_529217\|2019-12 | ···················· | ···················· | ···················· | ···················· | ···················· | ···················· | |
| A | 6657 | hCoV-19/Shenzhen/HKU-SZ-005/2020\|EPI_ISL_405839\|20 | ···················· | ···················· | ···················· | ···················· | ···················· | ···················· | |
| A | 6658 | hCoV-19/Shenzhen/HKU-SZ-002/2020\|EPI_ISL_406030\|20 | ···················· | ···················· | ···················· | ···················· | ···················· | ···················· | |
| A | 6659 | hCoV-19/USA/CA-CDC-1/2020\|EPI_ISL_406034\|2020-01-2 | ···················· | ···················· | ···················· | ···················· | ···················· | ···················· | |
| A | 6660 | hCoV-19/USA/CA2/2020\|EPI_ISL_406036\|2020-01-22 | ···················· | ···················· | ···················· | ···················· | ···················· | ···················· | |
| A | 6661 | hCoV-19/Japan/TY-WK-012/2020\|EPI_ISL_408665\|2020-0 | ···················· | ···················· | ···················· | ···················· | ···················· | ···················· | |
| A | 6662 | hCoV-19/Japan/TY-WK-501/2020\|EPI_ISL_408666\|2020-0 | ···················· | ···················· | ···················· | ···················· | ···················· | ···················· | |
| A | 6663 | hCoV-19/Japan/TY-WK-521/2020\|EPI_ISL_408667\|2020-0 | ···················· | ···················· | ···················· | ···················· | ···················· | ···················· | |
| A | 6664 | hCoV-19/Vietnam/VR03-38142/2020\|EPI_ISL_408668\|202 | ···················· | ···················· | ···················· | ···················· | ···················· | ···················· | |
| A | 6665 | hCoV-19/Japan/KY-V-029/2020\|EPI_ISL_408669\|2020-01 | ···················· | ···················· | ···················· | ···················· | ···················· | ···················· | |
| A | 6666 | hCoV-19/USA/WI1/2020\|EPI_ISL_408670\|2020-01-31 | ···················· | ···················· | ···················· | ···················· | ···················· | ···················· | |
| A | 6667 | hCoV-19/France/IDF-0515-isl/2020\|EPI_ISL_410984\|20 | ···················· | ···················· | ···················· | ···················· | ···················· | ···················· | |
| A | 6668 | hCoV-19/France/IDF-2279/2020\|EPI_ISL_416500\|2020-0 | ···················· | ···················· | ···················· | ···················· | ···················· | ···················· | |
| A | 6669 | hCoV-19/France/IDF-2284/2020\|EPI_ISL_416501\|2020-0 | ···················· | ···················· | ···················· | ···················· | ···················· | ···················· | |
| A | 6670 | hCoV-19/France/BRE-2330/2020\|EPI_ISL_416502\|2020-0 | ···················· | ···················· | ···················· | ···················· | ···················· | ···················· | |
| A | 6671 | hCoV-19/France/BRE-2334/2020\|EPI_ISL_416503\|2020-0 | ···················· | ···················· | ···················· | ···················· | ···················· | ···················· | |
| A | 6672 | hCoV-19/France/BRE-2335/2020\|EPI_ISL_416504\|2020-0 | ···················· | ···················· | ···················· | ···················· | ···················· | ···················· | |
| A | 6673 | hCoV-19/France/BRE-2336/2020\|EPI_ISL_416505\|2020-0 | ···················· | ···················· | ···················· | ···················· | ···················· | ···················· | |
| A | 6674 | hCoV-19/France/BRE-2337/2020\|EPI_ISL_416506\|2020-0 | ···················· | ···················· | ···················· | ···················· | ···················· | ···················· | |
| A | 6675 | hCoV-19/France/BRE-2340/2020\|EPI_ISL_416507\|2020-0 | ···················· | ···················· | ···················· | ···················· | ···················· | ···················· | |
| A | 6676 | hCoV-19/France/BRE-2343/2020\|EPI_ISL_416508\|2020-0 | ···················· | ···················· | ···················· | ···················· | ···················· | ···················· | |
| A | 6677 | hCoV-19/France/BRE-2344/2020\|EPI_ISL_416509\|2020-0 | ···················· | ···················· | ···················· | ···················· | ···················· | ···················· | |
| A | 6678 | hCoV-19/France/BRE-2346/2020\|EPI_ISL_416510\|2020-0 | ···················· | ···················· | ···················· | ···················· | ···················· | ···················· | |
| A | 6679 | hCoV-19/France/BRE-2348/2020\|EPI_ISL_416511\|2020-0 | ···················· | ···················· | ···················· | ···················· | ···················· | ···················· | |
| A | 6680 | hCoV-19/France/BRE-2349/2020\|EPI_ISL_416512\|2020-0 | ···················· | ···················· | ···················· | ···················· | ···················· | ···················· | |
| A | 6681 | hCoV-19/France/BRE-2351/2020\|EPI_ISL_416513\|2020-0 | ···················· | ···················· | ···················· | ···················· | ···················· | ···················· | |
| A | 6682 | hCoV-19/Australia/VIC08/2020\|EPI_ISL_416514\|2020-0 | ···················· | ···················· | ···················· | ···················· | ···················· | ···················· | |
| A | 6683 | hCoV-19/New Zealand/20VR0189/2020\|EPI_ISL_416519\|2 | ···················· | ···················· | ···················· | ···················· | ···················· | ···················· | |
| A | 6684 | hCoV-19/USA/WI-UW-03/2020\|EPI_ISL_416523\|2020-03-1 | ···················· | ···················· | ···················· | ···················· | ···················· | ···················· | |
| A | 6685 | hCoV-19/Japan/SMU-0311S2/2020\|EPI_ISL_416524\|2020- | ···················· | ···················· | ···················· | ···················· | ···················· | ···················· | |
| A | 6686 | hCoV-19/Japan/SMU-0311S3/2020\|EPI_ISL_416525\|2020- | ···················· | ···················· | ···················· | ···················· | ···················· | ···················· | |
| A | 6687 | hCoV-19/New Zealand/20VR0275/2020\|EPI_ISL_416538\|2 | ···················· | ···················· | ···················· | ···················· | ···················· | ···················· | |
| A | 6688 | hCoV-19/New Zealand/20VR0276/2020\|EPI_ISL_416539\|2 | ···················· | ···················· | ···················· | ···················· | ···················· | ···················· | |
| A | 6689 | hCoV-19/Kuwait/KU09/2020\|EPI_ISL_416541\|2020-03-02 | ···················· | ···················· | ···················· | ···················· | ···················· | ···················· | |
| A | 6690 | hCoV-19/Kuwait/KU17/2020\|EPI_ISL_416542\|2020-03-02 | ···················· | ···················· | ···················· | ···················· | ···················· | ···················· | |
| A | 6691 | hCoV-19/Kuwait/KU18/2020\|EPI_ISL_416543\|2020-03-02 | ···················· | ···················· | ···················· | ···················· | ···················· | ···················· | |
| A | 6692 | hCoV-19/Iceland/15/2020\|EPI_ISL_417800\|2020-03-02 | ···················· | ···················· | ···················· | ···················· | ···················· | ···················· | |
| A | 6693 | hCoV-19/Iceland/151/2020\|EPI_ISL_417801\|2020-03-13 | ···················· | ···················· | ···················· | ···················· | ···················· | ···················· | |
| A | 6694 | hCoV-19/Iceland/153/2020\|EPI_ISL_417802\|2020-03-13 | ···················· | ···················· | ···················· | ···················· | ···················· | ···················· | |
| A | 6695 | hCoV-19/Iceland/156/2020\|EPI_ISL_417803\|2020-03-13 | ···················· | ···················· | ···················· | ···················· | ···················· | ···················· | |
| A | 6696 | hCoV-19/Iceland/157/2020\|EPI_ISL_417804\|2020-03-13 | ···················· | ···················· | ···················· | ···················· | ···················· | ···················· | |
| A | 6697 | hCoV-19/Iceland/158/2020\|EPI_ISL_417805\|2020-03-13 | ···················· | ···················· | ···················· | ···················· | ···················· | ···················· | |
| A | 6698 | hCoV-19/Iceland/159/2020\|EPI_ISL_417806\|2020-03-13 | ···················· | ···················· | ···················· | ···················· | ···················· | ···················· | |
| A | 6699 | hCoV-19/Iceland/165/2020\|EPI_ISL_417808\|2020-03-15 | ···················· | ···················· | ···················· | ···················· | ···················· | ···················· | |
| A | 6700 | hCoV-19/Iceland/167/2020\|EPI_ISL_417809\|2020-03-15 | ···················· | ···················· | ···················· | ···················· | ···················· | ···················· | |
| A | 6701 | hCoV-19/Iceland/168/2020\|EPI_ISL_417810\|2020-03-13 | ···················· | ···················· | ···················· | ···················· | ···················· | ···················· | |
| A | 6702 | hCoV-19/Iceland/176/2020\|EPI_ISL_417812\|2020-03-14 | ···················· | ···················· | ···················· | ···················· | ···················· | ···················· | |
| A | 6703 | hCoV-19/Iceland/178/2020\|EPI_ISL_417813\|2020-03-15 | ···················· | ···················· | ···················· | ···················· | ···················· | ···················· | |
| A | 6704 | hCoV-19/Iceland/181/2020\|EPI_ISL_417814\|2020-03-16 | ···················· | ···················· | ···················· | ···················· | ···················· | ···················· | |
| A | 6705 | hCoV-19/Iceland/186/2020\|EPI_ISL_417815\|2020-03-16 | ···················· | ···················· | ···················· | ···················· | ···················· | ···················· | |
| A | 6706 | hCoV-19/Iceland/188/2020\|EPI_ISL_417816\|2020-03-16 | ···················· | ···················· | ···················· | ···················· | ···················· | ···················· | |
| A | 6707 | hCoV-19/Iceland/19/2020\|EPI_ISL_417817\|2020-03-02 | ···················· | ···················· | ···················· | ···················· | ···················· | ···················· | |
| A | 6708 | hCoV-19/Iceland/190/2020\|EPI_ISL_417818\|2020-03-16 | ···················· | ···················· | ···················· | ···················· | ···················· | ···················· | |

| | | | | | | | | | |
|---|---|---|---|---|---|---|---|---|---|
| A | 6709 hCoV-19/Iceland/192/2020\|EPI_ISL_417819\|2020-03-15 | ···················· | ···················· | ···················· | ···················· | ···················· | ···················· | | |
| A | 6710 hCoV-19/Iceland/193/2020\|EPI_ISL_417820\|2020-03-16 | ···················· | ···················· | ···················· | ···················· | ···················· | ···················· | | |
| A | 6711 hCoV-19/Iceland/196/2020\|EPI_ISL_417822\|2020-03-16 | ···················· | ···················· | ···················· | ···················· | ···················· | ···················· | | |
| A | 6712 hCoV-19/Iceland/198/2020\|EPI_ISL_417823\|2020-03-16 | ···················· | ···················· | ···················· | ···················· | ···················· | ···················· | | |
| A | 6713 hCoV-19/Iceland/201/2020\|EPI_ISL_417824\|2020-03-16 | ···················· | ···················· | ···················· | ···················· | ···················· | ···················· | | |
| A | 6714 hCoV-19/Iceland/203/2020\|EPI_ISL_417825\|2020-03-16 | ···················· | ···················· | ···················· | ···················· | ···················· | ···················· | | |
| A | 6715 hCoV-19/Iceland/204/2020\|EPI_ISL_417826\|2020-03-16 | ···················· | ···················· | ···················· | ···················· | ···················· | ···················· | | |
| A | 6716 hCoV-19/Iceland/205/2020\|EPI_ISL_417827\|2020-03-16 | ···················· | ···················· | ···················· | ···················· | ···················· | ···················· | | |
| A | 6718 hCoV-19/Iceland/210/2020\|EPI_ISL_417829\|2020-03-16 | ···················· | ···················· | ···················· | ···················· | ···················· | ···················· | | |
| A | 6719 hCoV-19/Iceland/212/2020\|EPI_ISL_417830\|2020-03-16 | ···················· | ···················· | ···················· | ···················· | ···················· | ···················· | | |
| A | 6720 hCoV-19/Iceland/214/2020\|EPI_ISL_417831\|2020-03-16 | ···················· | ···················· | ···················· | ···················· | ···················· | ···················· | | |
| A | 6721 hCoV-19/Iceland/216/2020\|EPI_ISL_417832\|2020-03-16 | ···················· | ···················· | ···················· | ···················· | ···················· | ···················· | | |
| A | 6722 hCoV-19/Iceland/217/2020\|EPI_ISL_417833\|2020-03-16 | ···················· | ···················· | ···················· | ···················· | ···················· | ···················· | | |
| A | 6723 hCoV-19/Iceland/218/2020\|EPI_ISL_417834\|2020-03-16 | ···················· | ···················· | ···················· | ···················· | ···················· | ···················· | | |
| A | 6724 hCoV-19/Iceland/219/2020\|EPI_ISL_417835\|2020-03-16 | ···················· | ···················· | ···················· | ···················· | ···················· | ···················· | | |
| A | 6725 hCoV-19/Iceland/221/2020\|EPI_ISL_417836\|2020-03-16 | ···················· | ···················· | ···················· | ···················· | ···················· | ···················· | | |
| A | 6726 hCoV-19/Iceland/222/2020\|EPI_ISL_417837\|2020-03-16 | ···················· | ···················· | ···················· | ···················· | ···················· | ···················· | | |
| A | 6727 hCoV-19/Iceland/223/2020\|EPI_ISL_417838\|2020-03-16 | ···················· | ···················· | ···················· | ···················· | ···················· | ···················· | | |
| A | 6728 hCoV-19/Iceland/224/2020\|EPI_ISL_417839\|2020-03-16 | ···················· | ···················· | ···················· | ···················· | ···················· | ···················· | | |
| A | 6729 hCoV-19/Iceland/23/2020\|EPI_ISL_417840\|2020-03-02 | ···················· | ···················· | ···················· | ···················· | ···················· | ···················· | | |
| A | 6730 hCoV-19/Iceland/26/2020\|EPI_ISL_417841\|2020-03-03 | ···················· | ···················· | ···················· | ···················· | ···················· | ···················· | | |
| A | 6731 hCoV-19/Iceland/31/2020\|EPI_ISL_417842\|2020-03-03 | ···················· | ···················· | ···················· | ···················· | ···················· | ···················· | | |
| A | 6733 hCoV-19/Iceland/36/2020\|EPI_ISL_417844\|2020-03-04 | ···················· | ···················· | ···················· | ···················· | ···················· | ···················· | | |
| A | 6734 hCoV-19/Iceland/4/2020\|EPI_ISL_417845\|2020-03-13 | ···················· | ···················· | ···················· | ···················· | ···················· | ···················· | | |
| A | 6735 hCoV-19/Iceland/40/2020\|EPI_ISL_417846\|2020-03-04 | ···················· | ···················· | ···················· | ···················· | ···················· | ···················· | | |
| A | 6737 hCoV-19/Iceland/52/2020\|EPI_ISL_417850\|2020-03-05 | ···················· | ···················· | ···················· | ···················· | ···················· | ·····NN··N·········· | | 1 |
| A | 6738 hCoV-19/Iceland/53/2020\|EPI_ISL_417851\|2020-03-05 | ···················· | ···················· | ···················· | ···················· | ···················· | ···················· | | |
| A | 6739 hCoV-19/Iceland/56/2020\|EPI_ISL_417852\|2020-03-06 | ···················· | ···················· | ···················· | ···················· | ···················· | ···················· | | |
| A | 6740 hCoV-19/Iceland/59/2020\|EPI_ISL_417853\|2020-03-06 | ···················· | ···················· | ···················· | ···················· | ···················· | ···················· | | |
| A | 6741 hCoV-19/Iceland/6/2020\|EPI_ISL_417854\|2020-03-13 | ···················· | ···················· | ···················· | ···················· | ···················· | ···················· | | |
| A | 6742 hCoV-19/Iceland/63/2020\|EPI_ISL_417855\|2020-03-07 | ···················· | ···················· | ···················· | ···················· | ···················· | ···················· | | |
| A | 6743 hCoV-19/Iceland/64/2020\|EPI_ISL_417856\|2020-03-07 | ···················· | ···················· | ···················· | ···················· | ···················· | ·················NNN | 1 | 1 |
| A | 6744 hCoV-19/Iceland/66/2020\|EPI_ISL_417857\|2020-03-07 | ···················· | ···················· | ···················· | ···················· | ···················· | ···················· | | |
| A | 6745 hCoV-19/Iceland/72/2020\|EPI_ISL_417860\|2020-03-08 | ···················· | ···················· | ···················· | ···················· | ···················· | ···················· | | |
| A | 6746 hCoV-19/Iceland/73/2020\|EPI_ISL_417861\|2020-03-09 | ···················· | ···················· | ···················· | ···················· | ···················· | ···················· | | |
| A | 6747 hCoV-19/Iceland/77/2020\|EPI_ISL_417862\|2020-03-09 | ···················· | ···················· | ···················· | ···················· | ···················· | ···················· | | |
| A | 6748 hCoV-19/Iceland/78/2020\|EPI_ISL_417863\|2020-03-09 | ···················· | ···················· | ···················· | ···················· | ···················· | ···················· | | |
| A | 6749 hCoV-19/Iceland/8/2020\|EPI_ISL_417864\|2020-03-15 | ···················· | ···················· | ···················· | ···················· | ···················· | ···················· | | |
| A | 6750 hCoV-19/Iceland/80/2020\|EPI_ISL_417865\|2020-03-09 | ···················· | ···················· | ···················· | ···················· | ···················· | ···················· | | |
| A | 6751 hCoV-19/Iceland/81/2020\|EPI_ISL_417866\|2020-03-09 | ···················· | ···················· | ···················· | ···················· | ···················· | ···················· | | |
| A | 6752 hCoV-19/Iceland/82/2020\|EPI_ISL_417867\|2020-03-09 | ···················· | ···················· | ···················· | ···················· | ···················· | ···················· | | |
| A | 6753 hCoV-19/Iceland/83/2020\|EPI_ISL_417868\|2020-03-09 | ···················· | ···················· | ···················· | ···················· | ···················· | ···················· | | |
| A | 6754 hCoV-19/Iceland/9/2020\|EPI_ISL_417871\|2020-03-11 | ···················· | ···················· | ···················· | ···················· | ···················· | ···················· | | |
| A | 6755 hCoV-19/Iceland/90/2020\|EPI_ISL_417872\|2020-03-10 | ···················· | ···················· | ···················· | ···················· | ···················· | ···················· | | |
| A | 6756 hCoV-19/Iceland/91/2020\|EPI_ISL_417873\|2020-03-10 | ···················· | ···················· | ···················· | ···················· | ···················· | ···················· | | |
| A | 6759 hCoV-19/Iceland/97/2020\|EPI_ISL_417876\|2020-03-10 | ···················· | ···················· | ···················· | ···················· | ···················· | ···················· | | |
| A | 6760 hCoV-19/Slovakia/SK-BMC1/2020\|EPI_ISL_417877\|2020- | ···················· | ···················· | ···················· | ···················· | ···················· | ···················· | | |
| A | 6761 hCoV-19/Slovakia/SK-BMC2/2020\|EPI_ISL_417878\|2020- | ···················· | ···················· | ···················· | ···················· | ···················· | ···················· | | |
| A | 6762 hCoV-19/Slovakia/SK-BMC5/2020\|EPI_ISL_417879\|2020- | ···················· | ···················· | ···················· | ···················· | ···················· | ···················· | | |
| A | 6763 hCoV-19/Slovakia/SK-BMC6/2020\|EPI_ISL_417880\|2020- | ···················· | ···················· | ···················· | ···················· | ···················· | ···················· | | |
| A | 6764 hCoV-19/Portugal/PT0025/2020\|EPI_ISL_418010\|2020-0 | ···················· | ···················· | ···················· | ···················· | ···················· | ···················· | | |
| A | 6765 hCoV-19/Portugal/PT0026/2020\|EPI_ISL_418011\|2020-0 | ···················· | ···················· | ···················· | ···················· | ···················· | ···················· | | |
| A | 6766 hCoV-19/Portugal/PT0027/2020\|EPI_ISL_418012\|2020-0 | ···················· | ···················· | ···················· | ···················· | ···················· | ···················· | | |
| A | 6767 hCoV-19/Portugal/PT0028/2020\|EPI_ISL_418013\|2020-0 | ···················· | ···················· | ···················· | ···················· | ···················· | ···················· | | |
| A | 6768 hCoV-19/Portugal/PT0029/2020\|EPI_ISL_418014\|2020-0 | ···················· | ···················· | ···················· | ···················· | ···················· | ···················· | | |
| A | 6769 hCoV-19/Portugal/PT0030/2020\|EPI_ISL_418015\|2020-0 | ···················· | ···················· | ···················· | ···················· | ···················· | ···················· | | |
| A | 6770 hCoV-19/Portugal/PT0031/2020\|EPI_ISL_418016\|2020-0 | ···················· | ···················· | ···················· | ···················· | ···················· | ···················· | | |
| A | 6771 hCoV-19/Portugal/PT0032/2020\|EPI_ISL_418017\|2020-0 | ···················· | ···················· | ···················· | ···················· | ···················· | ···················· | | |
| A | 6772 hCoV-19/Portugal/PT0033/2020\|EPI_ISL_418018\|2020-0 | ···················· | ···················· | ···················· | ···················· | ···················· | ···················· | | |
| A | 6773 hCoV-19/Portugal/PT0034/2020\|EPI_ISL_418019\|2020-0 | ···················· | ···················· | ···················· | ···················· | ···················· | ···················· | | |
| A | 6774 hCoV-19/Japan/P5-2/2020\|EPI_ISL_419310\|2020-03-13 | ···················· | ···················· | ···················· | ···················· | ···················· | ···················· | | |
| A | 6775 hCoV-19/Japan/P5-3/2020\|EPI_ISL_419311\|2020-03-13 | ···················· | ···················· | ···················· | ···················· | ···················· | ···················· | | |
| A | 6776 hCoV-19/USA/NY-NYUMC49/2020\|EPI_ISL_420300\|2020-03 | ···················· | ···················· | ···················· | ···················· | ···················· | ···················· | | |
| A | 6777 hCoV-19/USA/NY-NYUMC50/2020\|EPI_ISL_420301\|2020-03 | ···················· | ···················· | ···················· | ···················· | ···················· | ···················· | | |
| A | 6778 hCoV-19/USA/NY-NYUMC51/2020\|EPI_ISL_420302\|2020-03 | ···················· | ···················· | ···················· | ···················· | ···················· | ···················· | | |
| A | 6779 hCoV-19/USA/AK-PHL002/2020\|EPI_ISL_420303\|2020-03- | ···················· | ···················· | ···················· | ···················· | ···················· | ···················· | | |
| A | 6780 hCoV-19/USA/AK-PHL003/2020\|EPI_ISL_420304\|2020-03- | ···················· | ···················· | ···················· | ···················· | ···················· | ···················· | | |
| A | 6781 hCoV-19/USA/AK-PHL006/2020\|EPI_ISL_420305\|2020-03- | ···················· | ···················· | ···················· | ···················· | ···················· | ···················· | | |
| A | 6782 hCoV-19/USA/AK-PHL015/2020\|EPI_ISL_420306\|2020-03- | ···················· | ···················· | ···················· | ···················· | ···················· | ···················· | | |
| A | 6783 hCoV-19/USA/NY-NYUMC52/2020\|EPI_ISL_420307\|2020-03 | ···················· | ···················· | ···················· | ···················· | ···················· | ···················· | | |
| A | 6784 hCoV-19/USA/NY-NYUMC53/2020\|EPI_ISL_420308\|2020-03 | ···················· | ···················· | ···················· | ···················· | ···················· | ···················· | | |
| A | 6785 hCoV-19/USA/NY-NYUMC54/2020\|EPI_ISL_420309\|2020-03 | ···················· | ···················· | ···················· | ···················· | ···················· | ···················· | | |
| A | 6786 hCoV-19/Belgium/MCW-030574/2020\|EPI_ISL_420330\|202 | ···················· | ···················· | ···················· | ···················· | ···················· | ···················· | | |
| A | 6787 hCoV-19/Belgium/LY-030575/2020\|EPI_ISL_420331\|2020 | ···················· | ···················· | ···················· | ···················· | ···················· | ···················· | | |
| A | 6788 hCoV-19/Belgium/WAM-030676/2020\|EPI_ISL_420332\|202 | ···················· | ···················· | ···················· | ···················· | ···················· | ···················· | | |
| A | 6789 hCoV-19/Belgium/RS-030677/2020\|EPI_ISL_420333\|2020 | ···················· | ···················· | ···················· | ···················· | ···················· | ···················· | | |
| A | 6790 hCoV-19/Wales/PHWC-25524/2020\|EPI_ISL_421002\|2020- | ···················· | ····A··············· | ···················· | ···················· | ···················· | ···················· | | |
| A | 6791 hCoV-19/Wales/PHWC-25542/2020\|EPI_ISL_421003\|2020- | ···················· | ···················· | ···················· | ···················· | ···················· | ···················· | | |

| | | | | | | | | |
|---|---|---|---|---|---|---|---|---|
| A | 6792 | hCoV-19/Wales/PHWC-25551/2020\|EPI_ISL_421004\|2020- | ···················· | ···················· | ····················· | ···················· | ···················· | ···················· |
| A | 6793 | hCoV-19/Wales/PHWC-25560/2020\|EPI_ISL_421005\|2020- | ···················· | ···················· | ····················· | ···················· | ···················· | ···················· |
| A | 6794 | hCoV-19/Wales/PHWC-2557F/2020\|EPI_ISL_421006\|2020- | ···················· | ···················· | ····················· | ···················· | ···················· | ···················· |
| A | 6795 | hCoV-19/Wales/PHWC-255AC/2020\|EPI_ISL_421008\|2020- | ···················· | ···················· | ····················· | ···················· | ···················· | ···················· |
| A | 6796 | hCoV-19/Wales/PHWC-255D9/2020\|EPI_ISL_421009\|2020- | ···················· | ····A··············· | ····················· | ···················· | ···················· | ···················· |
| A | 6797 | hCoV-19/Wales/PHWC-255E8/2020\|EPI_ISL_421010\|2020- | ···················· | ····A··············· | ····················· | ···················· | ···················· | ···················· |
| A | 6798 | hCoV-19/Wales/PHWC-255F7/2020\|EPI_ISL_421011\|2020- | ···················· | ···················· | ····················· | ···················· | ···················· | ···················· |
| A | 6799 | hCoV-19/USA/NY-PV09075/2020\|EPI_ISL_421621\|2020-03 | ···················· | ···················· | ····················· | ···················· | ···················· | ···················· |
| A | 6800 | hCoV-19/USA/NY-PV09077/2020\|EPI_ISL_421623\|2020-03 | ···················· | ···················· | ····················· | ···················· | ···················· | ···················· |
| A | 6801 | hCoV-19/USA/NY-PV09079/2020\|EPI_ISL_421624\|2020-03 | ···················· | ···················· | ····················· | ···················· | ···················· | ···················· |
| A | 6802 | hCoV-19/USA/NY-PV09083/2020\|EPI_ISL_421626\|2020-03 | ···················· | ···················· | ····················· | ···················· | ···················· | ···················· |
| A | 6803 | hCoV-19/USA/NY-PV09087/2020\|EPI_ISL_421627\|2020-03 | ···················· | ···················· | ····················· | ···················· | ···················· | ···················· |
| A | 6804 | hCoV-19/USA/NY-PV09088/2020\|EPI_ISL_421628\|2020-03 | ···················· | ···················· | ····················· | ···················· | ···················· | ···················· |
| A | 6805 | hCoV-19/USA/NY-PV09090/2020\|EPI_ISL_421629\|2020-03 | ···················· | ···················· | ····················· | ···················· | ···················· | ···················· |
| A | 6806 | hCoV-19/USA/NY-PV09092/2020\|EPI_ISL_421630\|2020-03 | ···················· | ···················· | ····················· | ···················· | ···················· | ···················· |
| A | 6807 | hCoV-19/USA/NY-PV09097/2020\|EPI_ISL_421631\|2020-03 | ···················· | ···················· | ····················· | ···················· | ···················· | ···················· |
| A | 6808 | hCoV-19/USA/NY-PV09098/2020\|EPI_ISL_421632\|2020-03 | ···················· | ···················· | ····················· | ···················· | ···················· | ···················· |
| A | 6809 | hCoV-19/USA/NY-PV09099/2020\|EPI_ISL_421633\|2020-03 | ···················· | ···················· | ····················· | ···················· | ···················· | ···················· |
| A | 6810 | hCoV-19/USA/NY-PV09102/2020\|EPI_ISL_421634\|2020-03 | ···················· | ···················· | ····················· | ···················· | ···················· | ···················· |
| A | 6811 | hCoV-19/USA/NY-PV09103/2020\|EPI_ISL_421635\|2020-03 | ···················· | ···················· | ····················· | ···················· | ···················· | ···················· |
| A | 6812 | hCoV-19/Australia/NSWID930/2020\|EPI_ISL_421636\|202 | ···················· | ···················· | ····················· | ···················· | ···················· | ···················· |
| A | 6813 | hCoV-19/Taiwan/144/2020\|EPI_ISL_421641\|2020-03-19 | ···················· | ···················· | ····················· | ···················· | ···················· | ···················· |
| A | 6814 | hCoV-19/USA/OH-OSUP0109/2020\|EPI_ISL_421660\|2020-0 | ···················· | ···················· | ····················· | ···················· | ···················· | ···················· |
| A | 6815 | hCoV-19/India/UN-1073/2020\|EPI_ISL_421662\|2020-03- | ···················· | ···················· | ····················· | ···················· | ···················· | ···················· |
| A | 6816 | hCoV-19/India/UN-1093/2020\|EPI_ISL_421663\|2020-03- | ···················· | ···················· | ····················· | ···················· | ···················· | ···················· |
| A | 6817 | hCoV-19/India/UN-1100/2020\|EPI_ISL_421664\|2020-03- | ···················· | ···················· | ····················· | ···················· | ···················· | ···················· |
| A | 6818 | hCoV-19/Netherlands/ZuidHolland_84/2020\|EPI_ISL_42 | ···················· | ···················· | ····················· | ···················· | ···················· | ···················· |
| A | 6819 | hCoV-19/Netherlands/ZuidHolland_86/2020\|EPI_ISL_42 | ···················· | ···················· | ····················· | ···················· | ···················· | ···················· |
| A | 6820 | hCoV-19/Netherlands/ZuidHolland_88/2020\|EPI_ISL_42 | ···················· | ···················· | ····················· | ···················· | ···················· | ···················· |
| A | 6821 | hCoV-19/Netherlands/ZuidHolland_89/2020\|EPI_ISL_42 | ···················· | ···················· | ····················· | ···················· | ···················· | ···················· |
| A | 6822 | hCoV-19/Netherlands/ZuidHolland_90/2020\|EPI_ISL_42 | ···················· | ···················· | ····················· | ···················· | ···················· | ···················· |
| A | 6823 | hCoV-19/Netherlands/ZuidHolland_93/2020\|EPI_ISL_42 | ···················· | ···················· | ····················· | ···················· | ···················· | ···················· |
| A | 6824 | hCoV-19/USA/IL-UW-1265/2020\|EPI_ISL_422961\|2020-03 | ···················· | ···················· | ····················· | ···················· | ···················· | ···················· |
| A | 6825 | hCoV-19/USA/UN-UW-1312/2020\|EPI_ISL_422962\|2020-03 | ···················· | ···················· | ····················· | ···················· | ···················· | ···················· |
| A | 6826 | hCoV-19/USA/WA-UW-1271/2020\|EPI_ISL_422963\|2020-03 | ···················· | ···················· | ····················· | ···················· | ···················· | ···················· |
| A | 6827 | hCoV-19/USA/WA-UW-1304/2020\|EPI_ISL_422964\|2020-03 | ···················· | ···················· | ····················· | ···················· | ···················· | ···················· |
| A | 6828 | hCoV-19/USA/WA-UW-1315/2020\|EPI_ISL_422965\|2020-03 | ···················· | ···················· | ····················· | ···················· | ···················· | ···················· |
| A | 6829 | hCoV-19/USA/WA-UW-1329/2020\|EPI_ISL_422966\|2020-03 | ···················· | ···················· | ····················· | ···················· | ···················· | ···················· |
| A | 6830 | hCoV-19/USA/WA-UW-1302/2020\|EPI_ISL_422967\|2020-03 | ···················· | ···················· | ····················· | ···················· | ···················· | ···················· |
| A | 6831 | hCoV-19/USA/UN-UW-1331/2020\|EPI_ISL_422968\|2020-03 | ···················· | ···················· | ····················· | ···················· | ···················· | ···················· |
| A | 6832 | hCoV-19/USA/WA-UW-1287/2020\|EPI_ISL_422969\|2020-03 | ···················· | ···················· | ····················· | ···················· | ···················· | ···················· |
| A | 6833 | hCoV-19/USA/WA-UW-1269/2020\|EPI_ISL_422970\|2020-03 | ···················· | ···················· | ····················· | ···················· | ···················· | ···················· |
| A | 6834 | hCoV-19/USA/WA-UW-1320/2020\|EPI_ISL_422971\|2020-03 | ···················· | ···················· | ····················· | ···················· | ···················· | ···················· |
| A | 6835 | hCoV-19/USA/WA-UW-1319/2020\|EPI_ISL_422972\|2020-03 | ···················· | ···················· | ····················· | ···················· | ···················· | ···················· |
| A | 6836 | hCoV-19/USA/WA-UW-1308/2020\|EPI_ISL_422973\|2020-03 | ···················· | ···················· | ····················· | ···················· | ···················· | ···················· |
| A | 6837 | hCoV-19/USA/WA-UW-1295/2020\|EPI_ISL_422990\|2020-03 | ···················· | ···················· | ····················· | ···················· | ···················· | ···················· |
| A | 6838 | hCoV-19/USA/WA-UW-1314/2020\|EPI_ISL_422991\|2020-03 | ···················· | ···················· | ····················· | ···················· | ···················· | ···················· |
| A | 6839 | hCoV-19/USA/WA-UW-1347/2020\|EPI_ISL_422992\|2020-03 | ···················· | ···················· | ····················· | ···················· | ···················· | ···················· |
| A | 6840 | hCoV-19/USA/WA-UW-1321/2020\|EPI_ISL_422993\|2020-03 | ···················· | ···················· | ····················· | ···················· | ···················· | ···················· |
| A | 6841 | hCoV-19/USA/WA-UW-1322/2020\|EPI_ISL_422994\|2020-03 | ···················· | ···················· | ····················· | ···················· | ···················· | ···················· |
| A | 6842 | hCoV-19/USA/WA-UW-1334/2020\|EPI_ISL_422995\|2020-03 | ···················· | ···················· | ····················· | ···················· | ···················· | ···················· |
| A | 6843 | hCoV-19/England/20130063804/2020\|EPI_ISL_423648\|20 | ···················· | ···················· | ····················· | ···················· | ···················· | ···················· |
| A | 6845 | hCoV-19/England/20130065304/2020\|EPI_ISL_423654\|20 | ···················· | ···················· | ····················· | ···················· | ···················· | ···················· |
| A | 6846 | hCoV-19/England/20130072204/2020\|EPI_ISL_423665\|20 | ···················· | ···················· | ····················· | ···················· | ···················· | ···················· |
| A | 6849 | hCoV-19/England/20130092104/2020\|EPI_ISL_423679\|20 | ···················· | ···················· | ····················· | ···················· | ···················· | ···················· |
| A | 6850 | hCoV-19/England/20130092404/2020\|EPI_ISL_423682\|20 | ···················· | ···················· | ····················· | ···················· | ···················· | ···················· |
| A | 6852 | hCoV-19/Taiwan/CGMH-CGU-20/2020\|EPI_ISL_424978\|202 | ···················· | ···················· | ····················· | ···················· | ···················· | ···················· |
| A | 6853 | hCoV-19/USA/NC_200191/2020\|EPI_ISL_424983\|2020-03 | ···················· | ···················· | ····················· | ···················· | ···················· | ···················· |
| A | 6854 | hCoV-19/USA/NC_200181/2020\|EPI_ISL_424987\|2020-03 | ···················· | ···················· | ····················· | ···················· | ···················· | ···················· |
| A | 6855 | hCoV-19/France/OCC-KRA-ROB/2020\|EPI_ISL_424993\|202 | ···················· | ···················· | ····················· | ···················· | ···················· | ···················· |
| A | 6856 | hCoV-19/USA/WA-UW-3881/2020\|EPI_ISL_427200\|2020-03 | ···················· | ···················· | ····················· | ···················· | ···················· | ···················· |
| A | 6857 | hCoV-19/USA/WA-UW-3883/2020\|EPI_ISL_427201\|2020-03 | ···················· | ···················· | ····················· | ···················· | ···················· | ···················· |
| A | 6858 | hCoV-19/USA/WA-UW-3889/2020\|EPI_ISL_427202\|2020-03 | ···················· | ···················· | ····················· | ···················· | ···················· | ···················· |
| A | 6859 | hCoV-19/USA/WA-UW-3895/2020\|EPI_ISL_427203\|2020-03 | ···················· | ···················· | ····················· | ···················· | ···················· | ···················· |
| A | 6860 | hCoV-19/USA/WA-UW-3900/2020\|EPI_ISL_427204\|2020-03 | ···················· | ···················· | ····················· | ···················· | ···················· | ···················· |
| A | 6861 | hCoV-19/USA/WA-UW-3903/2020\|EPI_ISL_427205\|2020-03 | ···················· | ···················· | ····················· | ···················· | ···················· | ···················· |
| A | 6862 | hCoV-19/USA/WA-UW-3908/2020\|EPI_ISL_427206\|2020-03 | ···················· | ···················· | ····················· | ···················· | ···················· | ···················· |
| A | 6863 | hCoV-19/USA/WA-UW-3911/2020\|EPI_ISL_427207\|2020-03 | ···················· | ···················· | ····················· | ···················· | ···················· | ···················· |
| A | 6864 | hCoV-19/USA/WA-UW-3917/2020\|EPI_ISL_427208\|2020-03 | ···················· | ···················· | ····················· | ···················· | ···················· | ···················· |
| A | 6865 | hCoV-19/USA/WA-UW-4291/2020\|EPI_ISL_427255\|2020-03 | ···················· | ···················· | ····················· | ···················· | ···················· | ···················· |
| A | 6866 | hCoV-19/USA/WA-UW-4292/2020\|EPI_ISL_427256\|2020-03 | ···················· | ···················· | ····················· | ···················· | ···················· | ···················· |
| A | 6867 | hCoV-19/USA/WA-UW-4315/2020\|EPI_ISL_427257\|2020-03 | ···················· | ···················· | ····················· | ···················· | ···················· | ···················· |
| A | 6868 | hCoV-19/USA/WA-UW-4316/2020\|EPI_ISL_427258\|2020-03 | ···················· | ···················· | ····················· | ···················· | ···················· | ···················· |
| A | 6869 | hCoV-19/USA/WA-UW-4318/2020\|EPI_ISL_427259\|2020-03 | ···················· | ···················· | ····················· | ···················· | ···················· | ···················· |
| A | 6870 | hCoV-19/USA/WA-UW-4321/2020\|EPI_ISL_427260\|2020-03 | ···················· | ···················· | ····················· | ···················· | ···················· | ···················· |
| A | 6871 | hCoV-19/USA/WA-UW-4322/2020\|EPI_ISL_427261\|2020-03 | ···················· | ···················· | ····················· | ···················· | ···················· | ···················· |
| A | 6872 | hCoV-19/USA/WA-UW-4324/2020\|EPI_ISL_427262\|2020-03 | ···················· | ···················· | ····················· | ···················· | ···················· | ···················· |
| A | 6873 | hCoV-19/USA/WA-UW-4326/2020\|EPI_ISL_427263\|2020-03 | ···················· | ···················· | ····················· | ···················· | ···················· | ···················· |

| | | | | | | | | |
|---|---|---|---|---|---|---|---|---|
| A | 6874 | hCoV-19/USA/WA-UW-4329/2020\|EPI_ISL_427264\|2020-03 | ···················· | ···················· | ····················· | ···················· | ···················· | ···················· |
| A | 6875 | hCoV-19/USA/WA-UW-4330/2020\|EPI_ISL_427265\|2020-03 | ···················· | ···················· | ····················· | ···················· | ···················· | ···················· |
| A | 6876 | hCoV-19/USA/WA-UW-4335/2020\|EPI_ISL_427266\|2020-03 | ···················· | ···················· | ····················· | ···················· | ···················· | ···················· |
| A | 6877 | hCoV-19/USA/WA-UW-4340/2020\|EPI_ISL_427268\|2020-03 | ···················· | ···················· | ····················· | ···················· | ···················· | ···················· |
| A | 6878 | hCoV-19/USA/WA-UW-4499/2020\|EPI_ISL_427269\|2020-03 | ···················· | ···················· | ····················· | ···················· | ···················· | ···················· |
| A | 6879 | hCoV-19/Croatia/17-S7/2020\|EPI_ISL_429802\|2020-03 | ···················· | ···················· | ····················· | ···················· | ···················· | ···················· |
| A | 6880 | hCoV-19/Croatia/I7-S21/2020\|EPI_ISL_429803\|2020-03 | ···················· | ···················· | ····················· | ···················· | ···················· | ···················· |
| A | 6881 | hCoV-19/Croatia/7R-S8/2020\|EPI_ISL_429805\|2020-03 | ···················· | ···················· | ····················· | ···················· | ···················· | ···················· |
| A | 6882 | hCoV-19/Canada/NB_1/2020\|EPI_ISL_429806\|2020-03-09 | ···················· | ···················· | ····················· | ···················· | ···················· | ···················· |
| A | 6883 | hCoV-19/Canada/MB_2/2020\|EPI_ISL_429807\|2020-03-10 | ···················· | ···················· | ····················· | ···················· | ···················· | ···················· |
| A | 6884 | hCoV-19/Canada/NB_21/2020\|EPI_ISL_429811\|2020-03-1 | ···················· | ···················· | ····················· | ···················· | ···················· | ···················· |
| A | 6885 | hCoV-19/Canada/NS_30/2020\|EPI_ISL_429812\|2020-03-1 | ···················· | ···················· | ····················· | ···················· | ···················· | ···················· |
| A | 6886 | hCoV-19/Canada/NS_31/2020\|EPI_ISL_429813\|2020-03-1 | ···················· | ···················· | ····················· | ···················· | ···················· | ···················· |
| A | 6887 | hCoV-19/Canada/NS_32/2020\|EPI_ISL_429814\|2020-03-1 | ···················· | ···················· | ····················· | ···················· | ···················· | ···················· |
| A | 6888 | hCoV-19/Canada/NL_34/2020\|EPI_ISL_429815\|2020-03-1 | ···················· | ···················· | ····················· | ···················· | ···················· | ···················· |
| A | 6889 | hCoV-19/Canada/NB_35/2020\|EPI_ISL_429816\|2020-03-1 | ···················· | ···················· | ····················· | ···················· | ···················· | ···················· |
| A | 6890 | hCoV-19/Canada/MB_36/2020\|EPI_ISL_429817\|2020-03-1 | ···················· | ···················· | ····················· | ···················· | ···················· | ···················· |
| A | 6891 | hCoV-19/Canada/MB_37/2020\|EPI_ISL_429818\|2020-03-1 | ···················· | ···················· | ····················· | ···················· | ···················· | ···················· |
| A | 6892 | hCoV-19/Canada/MB_38/2020\|EPI_ISL_429819\|2020-03-1 | ···················· | ···················· | ····················· | ···················· | ···················· | ···················· |
| A | 6893 | hCoV-19/Canada/MB_39/2020\|EPI_ISL_429820\|2020-03-1 | ···················· | ···················· | ····················· | ···················· | ···················· | ···················· |
| A | 6894 | hCoV-19/USA/WI-GMF-00822/2020\|EPI_ISL_429843\|2020- | ···················· | ···················· | ····················· | ···················· | ···················· | ···················· |
| A | 6895 | hCoV-19/USA/WI-GMF-00857/2020\|EPI_ISL_429844\|2020- | ···················· | ···················· | ····················· | ···················· | ···················· | ···················· |
| A | 6896 | hCoV-19/USA/WI-GMF-01047/2020\|EPI_ISL_429845\|2020- | ···················· | ···················· | ····················· | ···················· | ···················· | ···················· |
| A | 6897 | hCoV-19/USA/WI-GMF-01158/2020\|EPI_ISL_429846\|2020- | ···················· | ···················· | ····················· | ···················· | ···················· | ···················· |
| A | 6898 | hCoV-19/USA/WI-GMF-01159/2020\|EPI_ISL_429847\|2020- | ···················· | ···················· | ····················· | ···················· | ···················· | ···················· |
| A | 6899 | hCoV-19/USA/WI-GMF-01316/2020\|EPI_ISL_429848\|2020- | ···················· | ···················· | ····················· | ···················· | ···················· | ···················· |
| A | 6900 | hCoV-19/Lishui/LS111/2020\|EPI_ISL_429852\|2020-01-2 | ···················· | ···················· | ····················· | ···················· | ···················· | ···················· |
| A | 6901 | hCoV-19/Lishui/LS556/2020\|EPI_ISL_429853\|2020-01-2 | ···················· | ···················· | ····················· | ···················· | ···················· | ···················· |
| A | 6902 | hCoV-19/Lishui/LS557/2020\|EPI_ISL_429854\|2020-02-0 | ···················· | ···················· | ····················· | ···················· | ···················· | ···················· |
| A | 6903 | hCoV-19/Turkey/HSGM-8968/2020\|EPI_ISL_429861\|2020- | ···················· | ·A·················· | ····················· | ···················· | ···················· | ··A················· |
| A | 6904 | hCoV-19/Turkey/HSGM-8970/2020\|EPI_ISL_429862\|2020- | ···················· | ···················· | ····················· | ···················· | ···················· | ···················· |
| A | 6905 | hCoV-19/Turkey/HSGM-8975/2020\|EPI_ISL_429863\|2020- | ···················· | ·A·················· | ····················· | ···················· | ···················· | ··A················· |
| A | 6906 | hCoV-19/Turkey/HSGM-8979/2020\|EPI_ISL_429864\|2020- | ···················· | ···················· | ····················· | ···················· | ···················· | ···················· |
| A | 6907 | hCoV-19/Turkey/HSGM-8992/2020\|EPI_ISL_429865\|2020- | ···················· | ···················· | ····················· | ···················· | ···················· | ···················· |
| A | 6908 | hCoV-19/Turkey/HSGM-4236/2020\|EPI_ISL_429866\|2020- | ···················· | ···················· | ····················· | ···················· | ···················· | ···················· |
| A | 6909 | hCoV-19/Turkey/HSGM-4505/2020\|EPI_ISL_429867\|2020- | ···················· | ···················· | ····················· | ···················· | ···················· | ···················· |
| A | 6910 | hCoV-19/Turkey/HSGM-4598/2020\|EPI_ISL_429868\|2020- | ···················· | ···················· | ····················· | ···················· | ···················· | ···················· |
| A | 6911 | hCoV-19/Turkey/HSGM-4701/2020\|EPI_ISL_429869\|2020- | ···················· | ·A·················· | ····················· | ···················· | ···················· | ··A················· |
| A | 6912 | hCoV-19/Germany/BY-ChVir-1019/2020\|EPI_ISL_450199\| | ···················· | ···················· | ····················· | ···················· | ···················· | ···················· |
| A | 6913 | hCoV-19/Taiwan/2/2020\|EPI_ISL_406031\|2020-01-23 | ···················· | ···················· | ····················· | ···················· | ···················· | ···················· |
| A | 6914 | hCoV-19/Portugal/PT0015/2020\|EPI_ISL_418000\|2020-0 | ···················· | ···················· | ····················· | ···················· | ···················· | ···················· |
| A | 6915 | hCoV-19/Portugal/PT0016/2020\|EPI_ISL_418001\|2020-0 | ···················· | ···················· | ····················· | ···················· | ···················· | ···················· |
| A | 6916 | hCoV-19/Portugal/PT0017/2020\|EPI_ISL_418002\|2020-0 | ···················· | ···················· | ····················· | ···················· | ···················· | ···················· |
| A | 6917 | hCoV-19/Portugal/PT0018/2020\|EPI_ISL_418003\|2020-0 | ···················· | ···················· | ····················· | ···················· | ···················· | ···················· |
| A | 6918 | hCoV-19/Portugal/PT0019/2020\|EPI_ISL_418004\|2020-0 | ···················· | ···················· | ····················· | ···················· | ···················· | ···················· |
| A | 6919 | hCoV-19/Portugal/PT0020/2020\|EPI_ISL_418005\|2020-0 | ···················· | ···················· | ····················· | ···················· | ···················· | ···················· |
| A | 6920 | hCoV-19/Portugal/PT0021/2020\|EPI_ISL_418006\|2020-0 | ···················· | ···················· | ····················· | ···················· | ···················· | ···················· |
| A | 6921 | hCoV-19/Portugal/PT0022/2020\|EPI_ISL_418007\|2020-0 | ···················· | ···················· | ····················· | ···················· | ···················· | ···················· |
| A | 6922 | hCoV-19/Portugal/PT0023/2020\|EPI_ISL_418008\|2020-0 | ···················· | ···················· | ····················· | ···················· | ···················· | ···················· |
| A | 6923 | hCoV-19/Portugal/PT0024/2020\|EPI_ISL_418009\|2020-0 | ···················· | ···················· | ····················· | ···················· | ···················· | ···················· |
| A | 6924 | hCoV-19/Portugal/PT0035/2020\|EPI_ISL_418020\|2020-0 | ···················· | ···················· | ····················· | ···················· | ···················· | ···················· |
| A | 6925 | hCoV-19/Portugal/PT0036/2020\|EPI_ISL_418021\|2020-0 | ···················· | ···················· | ····················· | ···················· | ···················· | ···················· |
| A | 6926 | hCoV-19/Portugal/PT0037/2020\|EPI_ISL_418022\|2020-0 | ···················· | ···················· | ····················· | ···················· | ···················· | ···················· |
| A | 6927 | hCoV-19/Portugal/PT0038/2020\|EPI_ISL_418023\|2020-0 | ···················· | ···················· | ····················· | ···················· | ···················· | ···················· |
| A | 6928 | hCoV-19/Portugal/PT0039/2020\|EPI_ISL_418024\|2020-0 | ···················· | ···················· | ····················· | ···················· | ···················· | ···················· |
| A | 6929 | hCoV-19/Portugal/PT0040/2020\|EPI_ISL_418025\|2020-0 | ···················· | ···················· | ····················· | ···················· | ···················· | ···················· |
| A | 6930 | hCoV-19/Portugal/PT0041/2020\|EPI_ISL_418026\|2020-0 | ···················· | ···················· | ····················· | ···················· | ···················· | ···················· |
| A | 6931 | hCoV-19/Portugal/PT0042/2020\|EPI_ISL_418027\|2020-0 | ···················· | ···················· | ····················· | ···················· | ···················· | ···················· |
| A | 6932 | hCoV-19/USA/MN-UW243/2020\|EPI_ISL_418028\|2020-03-1 | ···················· | ···················· | ····················· | ···················· | ···················· | ···················· |
| A | 6933 | hCoV-19/USA/WA-UW244/2020\|EPI_ISL_418029\|2020-03-1 | ···················· | ···················· | ····················· | ···················· | ···················· | ···················· |
| A | 6934 | hCoV-19/USA/WA-UW245/2020\|EPI_ISL_418030\|2020-03-1 | ···················· | ···················· | ····················· | ···················· | ···················· | ···················· |
| A | 6935 | hCoV-19/USA/WA-UW247/2020\|EPI_ISL_418032\|2020-03-1 | ···················· | ···················· | ····················· | ···················· | ···················· | ···················· |
| A | 6936 | hCoV-19/USA/WA-UW248/2020\|EPI_ISL_418033\|2020-03-1 | ···················· | ···················· | ····················· | ···················· | ···················· | ···················· |
| A | 6937 | hCoV-19/USA/WA-UW249/2020\|EPI_ISL_418034\|2020-03-1 | ···················· | ···················· | ····················· | ···················· | ···················· | ···················· |
| A | 6938 | hCoV-19/USA/MN-UW251/2020\|EPI_ISL_418036\|2020-03-1 | ···················· | ···················· | ····················· | ···················· | ···················· | ···················· |
| A | 6939 | hCoV-19/USA/WA-UW252/2020\|EPI_ISL_418037\|2020-03-1 | ···················· | ···················· | ····················· | ···················· | ···················· | ···················· |
| A | 6940 | hCoV-19/USA/WA-UW253/2020\|EPI_ISL_418038\|2020-03-1 | ···················· | ···················· | ····················· | ···················· | ···················· | ···················· |
| A | 6941 | hCoV-19/USA/CT-UW254/2020\|EPI_ISL_418039\|2020-03-1 | ···················· | ···················· | ····················· | ···················· | ···················· | ···················· |
| A | 6942 | hCoV-19/USA/WA-UW255/2020\|EPI_ISL_418040\|2020-03-1 | ···················· | ···················· | ····················· | ···················· | ···················· | ···················· |
| A | 6943 | hCoV-19/USA/CT-UW256/2020\|EPI_ISL_418041\|2020-03-1 | ···················· | ···················· | ····················· | ···················· | ···················· | ···················· |
| A | 6944 | hCoV-19/USA/MN-UW257/2020\|EPI_ISL_418042\|2020-03-1 | ···················· | ···················· | ····················· | ···················· | ···················· | ···················· |
| A | 6945 | hCoV-19/USA/CT-UW258/2020\|EPI_ISL_418043\|2020-03-1 | ···················· | ···················· | ····················· | ···················· | ···················· | ···················· |
| A | 6946 | hCoV-19/USA/CT-UW259/2020\|EPI_ISL_418044\|2020-03-1 | ···················· | ···················· | ····················· | ···················· | ···················· | ···················· |
| A | 6947 | hCoV-19/USA/CT-UW260/2020\|EPI_ISL_418045\|2020-03-1 | ···················· | ···················· | ····················· | ···················· | ···················· | ···················· |
| A | 6948 | hCoV-19/USA/WA-UW261/2020\|EPI_ISL_418046\|2020-03-1 | ···················· | ···················· | ····················· | ···················· | ···················· | ···················· |
| A | 6949 | hCoV-19/USA/WA-UW262/2020\|EPI_ISL_418047\|2020-03-1 | ···················· | ···················· | ····················· | ···················· | ···················· | ···················· |
| A | 6950 | hCoV-19/USA/MN-UW264/2020\|EPI_ISL_418049\|2020-03-1 | ···················· | ···················· | ····················· | ···················· | ···················· | ···················· |
| A | 6951 | hCoV-19/USA/WA-UW265/2020\|EPI_ISL_418050\|2020-03-1 | ···················· | ···················· | ····················· | ···················· | ···················· | ···················· |

| | | | | | | | | |
|---|---|---|---|---|---|---|---|---|
| A | 6952 | hCoV-19/USA/MN-UW266/2020\|EPI_ISL_418051\|2020-03-1 | .................... | .................... | .................... | .................... | .................... | .................... |
| A | 6953 | hCoV-19/USA/WA-UW268/2020\|EPI_ISL_418053\|2020-03-1 | .................... | .................... | .................... | .................... | .................... | .................... |
| A | 6954 | hCoV-19/USA/WA-UW269/2020\|EPI_ISL_418054\|2020-03-1 | .................... | .................... | .................... | .................... | .................... | .................... |
| A | 6955 | hCoV-19/USA/WA-UW270/2020\|EPI_ISL_418055\|2020-03-1 | .................... | .................... | .................... | .................... | .................... | .................... |
| A | 6956 | hCoV-19/USA/CT-UW271/2020\|EPI_ISL_418056\|2020-03-1 | .................... | .................... | .................... | .................... | .................... | .................... |
| A | 6957 | hCoV-19/USA/CT-UW272/2020\|EPI_ISL_418057\|2020-03-1 | .................... | .................... | .................... | .................... | .................... | .................... |
| A | 6958 | hCoV-19/USA/CT-UW273/2020\|EPI_ISL_418058\|2020-03-1 | .................... | .................... | .................... | .................... | .................... | .................... |
| A | 6959 | hCoV-19/USA/WA-UW274/2020\|EPI_ISL_418059\|2020-03-1 | .................... | .................... | .................... | .................... | .................... | .................... |
| A | 6960 | hCoV-19/Japan/P3-2/2020\|EPI_ISL_419300\|2020-03-20 | .................... | .................... | .................... | .................... | .................... | .................... |
| A | 6961 | hCoV-19/Japan/P4-1/2020\|EPI_ISL_419301\|2020-03-11 | .................... | .................... | .................... | .................... | .................... | .................... |
| A | 6962 | hCoV-19/Japan/P4-2/2020\|EPI_ISL_419302\|2020-03-12 | .................... | .................... | .................... | .................... | .................... | .................... |
| A | 6963 | hCoV-19/Japan/P4-3/2020\|EPI_ISL_419303\|2020-03-12 | .................... | .................... | .................... | .................... | .................... | .................... |
| A | 6964 | hCoV-19/Japan/P4-4/2020\|EPI_ISL_419304\|2020-03-17 | .................... | .................... | .................... | .................... | .................... | .................... |
| A | 6965 | hCoV-19/Japan/P4-5/2020\|EPI_ISL_419305\|2020-03-18 | .................... | .................... | .................... | .................... | .................... | .................... |
| A | 6966 | hCoV-19/Japan/P4-6/2020\|EPI_ISL_419306\|2020-03-20 | .................... | .................... | .................... | .................... | .................... | .................... |
| A | 6967 | hCoV-19/Japan/P4-7/2020\|EPI_ISL_419307\|2020-03-20 | .................... | .................... | .................... | .................... | .................... | .................... |
| A | 6968 | hCoV-19/Japan/P4-8/2020\|EPI_ISL_419308\|2020-03-19 | .................... | .................... | .................... | .................... | .................... | .................... |
| A | 6969 | hCoV-19/Japan/P5-1/2020\|EPI_ISL_419309\|2020-03-12 | .................... | .................... | .................... | .................... | .................... | .................... |
| A | 6970 | hCoV-19/Portugal/IGC00009/2020\|EPI_ISL_419386\|2020 | .................... | .................... | .................... | .................... | .................... | .................... |
| A | 6971 | hCoV-19/Portugal/IGC00010/2020\|EPI_ISL_419387\|2020 | .................... | .................... | .................... | .................... | .................... | .................... |
| A | 6972 | hCoV-19/USA/MN60-MDH60/2020\|EPI_ISL_419388\|2020-03 | .................... | .................... | .................... | .................... | .................... | .................... |
| A | 6973 | hCoV-19/Norway/2114/2020\|EPI_ISL_420312\|2020-03-18 | .................... | .................... | .................... | .................... | .................... | .................... |
| A | 6974 | hCoV-19/Norway/1953/2020\|EPI_ISL_420313\|2020-03-10 | .................... | .................... | .................... | .................... | .................... | .................... |
| A | 6975 | hCoV-19/Belgium/NL-030447/2020\|EPI_ISL_420314\|2020 | .................... | .................... | .................... | .................... | .................... | .................... |
| A | 6976 | hCoV-19/Belgium/RYR-030649/2020\|EPI_ISL_420315\|202 | .................... | .................... | .................... | .................... | .................... | .................... |
| A | 6977 | hCoV-19/Belgium/QOJ-030751/2020\|EPI_ISL_420316\|202 | .................... | .................... | .................... | .................... | .................... | .................... |
| A | 6978 | hCoV-19/Belgium/RJ-030552/2020\|EPI_ISL_420317\|2020 | .................... | .................... | .................... | .................... | .................... | .................... |
| A | 6979 | hCoV-19/Belgium/TG-030757/2020\|EPI_ISL_420318\|2020 | .................... | .................... | .................... | .................... | .................... | .................... |
| A | 6980 | hCoV-19/Belgium/GJ-030458/2020\|EPI_ISL_420319\|2020 | .................... | .................... | .................... | .................... | .................... | .................... |
| A | 6981 | hCoV-19/Belgium/030959/2020\|EPI_ISL_420320\|2020-03 | .................... | .................... | .................... | .................... | .................... | .................... |
| A | 6982 | hCoV-19/Belgium/WWM-030665/2020\|EPI_ISL_420321\|202 | .................... | .................... | .................... | .................... | .................... | .................... |
| A | 6983 | hCoV-19/Belgium/DBOM-030566/2020\|EPI_ISL_420322\|20 | .................... | .................... | .................... | .................... | .................... | .................... |
| A | 6984 | hCoV-19/Belgium/BJ-030767/2020\|EPI_ISL_420323\|2020 | .................... | .................... | .................... | .................... | .................... | .................... |
| A | 6985 | hCoV-19/Belgium/DVBJ-030468/2020\|EPI_ISL_420324\|20 | .................... | .................... | .................... | .................... | .................... | .................... |
| A | 6986 | hCoV-19/Belgium/DLDJ-030569/2020\|EPI_ISL_420325\|20 | .................... | .................... | .................... | .................... | .................... | .................... |
| A | 6987 | hCoV-19/Belgium/AKM-030670/2020\|EPI_ISL_420326\|202 | .................... | .................... | .................... | .................... | .................... | .................... |
| A | 6988 | hCoV-19/Belgium/HL-030771/2020\|EPI_ISL_420327\|2020 | .................... | .................... | .................... | .................... | .................... | .................... |
| A | 6989 | hCoV-19/Belgium/VGA-030672/2020\|EPI_ISL_420328\|202 | .................... | .................... | .................... | .................... | .................... | .................... |
| A | 6990 | hCoV-19/Belgium/GE-030573/2020\|EPI_ISL_420329\|2020 | .................... | .................... | .................... | .................... | .................... | .................... |
| A | 6991 | hCoV-19/Belgium/GOF-030578/2020\|EPI_ISL_420334\|202 | .................... | .................... | .................... | .................... | .................... | .................... |
| A | 6992 | hCoV-19/Belgium/CD-030679/2020\|EPI_ISL_420335\|2020 | .................... | .................... | .................... | .................... | .................... | .................... |
| A | 6993 | hCoV-19/Belgium/PA-030680/2020\|EPI_ISL_420336\|2020 | .................... | .................... | .................... | .................... | .................... | .................... |
| A | 6994 | hCoV-19/Belgium/PAN-030681/2020\|EPI_ISL_420337\|202 | .................... | .................... | .................... | .................... | .................... | .................... |
| A | 6995 | hCoV-19/Belgium/AC-030982/2020\|EPI_ISL_420338\|2020 | .................... | .................... | .................... | .................... | .................... | .................... |
| A | 6996 | hCoV-19/Belgium/SJ-030583/2020\|EPI_ISL_420339\|2020 | .................... | .................... | .................... | .................... | .................... | .................... |
| A | 6997 | hCoV-19/Belgium/MJP-030684/2020\|EPI_ISL_420340\|202 | .................... | .................... | .................... | .................... | .................... | .................... |
| A | 6998 | hCoV-19/Belgium/LM-030685/2020\|EPI_ISL_420341\|2020 | .................... | .................... | .................... | .................... | .................... | .................... |
| A | 6999 | hCoV-19/Belgium/ECC-030686/2020\|EPI_ISL_420342\|202 | .................... | .................... | .................... | .................... | .................... | .................... |
| A | 7000 | hCoV-19/Belgium/BM-030687/2020\|EPI_ISL_420343\|2020 | .................... | .................... | .................... | .................... | .................... | .................... |
| A | 7001 | hCoV-19/Belgium/RJL-030588/2020\|EPI_ISL_420344\|202 | .................... | .................... | .................... | .................... | .................... | .................... |
| A | 7002 | hCoV-19/Belgium/MC-030689/2020\|EPI_ISL_420345\|2020 | .................... | .................... | .................... | .................... | .................... | .................... |
| A | 7003 | hCoV-19/Belgium/SB-030990/2020\|EPI_ISL_420346\|2020 | .................... | .................... | .................... | .................... | .................... | .................... |
| A | 7004 | hCoV-19/Belgium/DA-030691/2020\|EPI_ISL_420347\|2020 | .................... | .................... | .................... | .................... | .................... | .................... |
| A | 7005 | hCoV-19/Belgium/FM-030592/2020\|EPI_ISL_420348\|2020 | .................... | .................... | .................... | .................... | .................... | .................... |
| A | 7006 | hCoV-19/Belgium/DD-030593/2020\|EPI_ISL_420349\|2020 | .................... | .................... | .................... | .................... | .................... | .................... |
| A | 7007 | hCoV-19/Belgium/BDW-030694/2020\|EPI_ISL_420350\|202 | .................... | .................... | .................... | .................... | .................... | .................... |
| A | 7008 | hCoV-19/Belgium/VMC-030695/2020\|EPI_ISL_420351\|202 | .................... | .................... | .................... | .................... | .................... | .................... |
| A | 7009 | hCoV-19/Belgium/DCS-030796/2020\|EPI_ISL_420352\|202 | .................... | .................... | .................... | .................... | .................... | .................... |
| A | 7010 | hCoV-19/Belgium/LD-030597/2020\|EPI_ISL_420353\|2020 | .................... | .................... | .................... | .................... | .................... | .................... |
| A | 7011 | hCoV-19/Belgium/BS-030598/2020\|EPI_ISL_420354\|2020 | .................... | .................... | .................... | .................... | .................... | .................... |
| A | 7012 | hCoV-19/Belgium/RR-030699/2020\|EPI_ISL_420355\|2020 | .................... | .................... | .................... | .................... | .................... | .................... |
| A | 7013 | hCoV-19/Belgium/MJP-0306100/2020\|EPI_ISL_420356\|20 | .................... | .................... | .................... | .................... | .................... | .................... |
| A | 7014 | hCoV-19/Belgium/JLJ-0320101/2020\|EPI_ISL_420357\|20 | .................... | .................... | .................... | .................... | .................... | .................... |
| A | 7015 | hCoV-19/Belgium/VD-0318102/2020\|EPI_ISL_420358\|202 | .................... | .................... | .................... | .................... | .................... | .................... |
| A | 7016 | hCoV-19/Belgium/UM-0318103/2020\|EPI_ISL_420359\|202 | .................... | .................... | .................... | .................... | .................... | .................... |
| A | 7017 | hCoV-19/Belgium/VL-0318104/2020\|EPI_ISL_420360\|202 | .................... | .................... | .................... | .................... | .................... | .................... |
| A | 7018 | hCoV-19/Belgium/GE-0317105/2020\|EPI_ISL_420361\|202 | .................... | .................... | .................... | .................... | .................... | .................... |
| A | 7019 | hCoV-19/Belgium/DA-0319106/2020\|EPI_ISL_420362\|202 | .................... | .................... | .................... | .................... | .................... | .................... |
| A | 7020 | hCoV-19/Belgium/VNP-0320107/2020\|EPI_ISL_420363\|20 | .................... | .................... | .................... | .................... | .................... | .................... |
| A | 7021 | hCoV-19/Belgium/HP-0319108/2020\|EPI_ISL_420364\|202 | .................... | .................... | .................... | .................... | .................... | .................... |
| A | 7022 | hCoV-19/Belgium/OT-0319109/2020\|EPI_ISL_420365\|202 | .................... | .................... | .................... | .................... | .................... | .................... |
| A | 7023 | hCoV-19/Belgium/KKN-0319184/2020\|EPI_ISL_420366\|20 | .................... | .................... | .................... | .................... | .................... | .................... |
| A | 7024 | hCoV-19/Belgium/JK-0305110/2020\|EPI_ISL_420367\|202 | .................... | .................... | .................... | .................... | .................... | .................... |
| A | 7025 | hCoV-19/Belgium/CA-0319111/2020\|EPI_ISL_420368\|202 | .................... | .................... | .................... | .................... | .................... | .................... |
| A | 7026 | hCoV-19/Belgium/SR-0319112/2020\|EPI_ISL_420369\|202 | .................... | .................... | .................... | .................... | .................... | .................... |
| A | 7027 | hCoV-19/Belgium/SC-0319113/2020\|EPI_ISL_420370\|202 | .................... | .................... | .................... | .................... | .................... | .................... |
| A | 7028 | hCoV-19/Belgium/VRJM-0323114/2020\|EPI_ISL_420371\|2 | .................... | .................... | .................... | .................... | .................... | .................... |
| A | 7029 | hCoV-19/Belgium/LPDC-0323115/2020\|EPI_ISL_420372\|2 | .................... | .................... | .................... | .................... | .................... | .................... |

| | | | | | | | | |
|---|---|---|---|---|---|---|---|---|
| A | 7030 | hCoV-19/Belgium/EBN-0323116/2020\|EPI_ISL_420373\|20 | ···················· | ···················· | ····················· | ···················· | ···················· | ···················· |
| A | 7031 | hCoV-19/Belgium/DV-0324117/2020\|EPI_ISL_420374\|202 | ···················· | ···················· | ····················· | ···················· | ···················· | ···················· |
| A | 7032 | hCoV-19/Belgium/VHV-0324118/2020\|EPI_ISL_420375\|20 | ···················· | ···················· | ····················· | ···················· | ···················· | ···················· |
| A | 7033 | hCoV-19/Belgium/CF-0324119/2020\|EPI_ISL_420376\|202 | ···················· | ···················· | ····················· | ···················· | ···················· | ···················· |
| A | 7034 | hCoV-19/Belgium/SEM-0324120/2020\|EPI_ISL_420377\|20 | ···················· | ···················· | ····················· | ···················· | ···················· | ···················· |
| A | 7035 | hCoV-19/USA/NY-PV09044/2020\|EPI_ISL_421607\|2020-03 | ···················· | ···················· | ····················· | ···················· | ···················· | ···················· |
| A | 7036 | hCoV-19/USA/NY-PV09056/2020\|EPI_ISL_421611\|2020-03 | ···················· | ···················· | ····················· | ···················· | ···················· | ···················· |
| A | 7037 | hCoV-19/USA/NY-PV09060/2020\|EPI_ISL_421612\|2020-03 | ···················· | ···················· | ····················· | ···················· | ···················· | ···················· |
| A | 7038 | hCoV-19/USA/NY-PV09061/2020\|EPI_ISL_421613\|2020-03 | ···T················ | ···················· | ····················· | ···················· | ···················· | ···················· |
| A | 7039 | hCoV-19/USA/NY-PV09063/2020\|EPI_ISL_421614\|2020-03 | ···················· | ···················· | ····················· | ···················· | ···················· | ···················· |
| A | 7040 | hCoV-19/USA/NY-PV09064/2020\|EPI_ISL_421615\|2020-03 | ·T·················· | ···················· | ····················· | ···················· | ···················· | ···················· |
| A | 7041 | hCoV-19/USA/NY-PV09067/2020\|EPI_ISL_421616\|2020-03 | ···················· | ···················· | ····················· | ···················· | ···················· | ···················· |
| A | 7042 | hCoV-19/USA/NY-PV09068/2020\|EPI_ISL_421617\|2020-03 | ···················· | ···················· | ····················· | ···················· | ···················· | ···················· |
| A | 7043 | hCoV-19/USA/NJ-PV09072/2020\|EPI_ISL_421619\|2020-03 | ···················· | ···················· | ····················· | ···················· | ···················· | ···················· |
| A | 7044 | hCoV-19/USA/NY-PV09073/2020\|EPI_ISL_421620\|2020-03 | ···················· | ···················· | ····················· | ···················· | ···················· | ···················· |
| A | 7045 | hCoV-19/Taiwan/225/2020\|EPI_ISL_421651\|2020-03-22 | ···················· | ···················· | ····················· | ···················· | ···················· | ···················· |
| A | 7046 | hCoV-19/Kuwait/KU001/2020\|EPI_ISL_421652\|2020-03-1 | ···················· | ···················· | ····················· | ···················· | ···················· | ···················· |
| A | 7047 | hCoV-19/Latvia/01/2020\|EPI_ISL_421653\|2020-03-25 | ···················· | ···················· | ····················· | ···················· | ···················· | ···················· |
| A | 7048 | hCoV-19/Latvia/02/2020\|EPI_ISL_421654\|2020-03-25 | ···················· | ···················· | ····················· | ···················· | ···················· | ···················· |
| A | 7049 | hCoV-19/Latvia/03/2020\|EPI_ISL_421655\|2020-03-25 | ···················· | ···················· | ····················· | ···················· | ···················· | ···················· |
| A | 7050 | hCoV-19/Latvia/04/2020\|EPI_ISL_421656\|2020-03-25 | ···················· | ···················· | ····················· | ···················· | ···················· | ···················· |
| A | 7051 | hCoV-19/India/UN-1104/2020\|EPI_ISL_421665\|2020-03- | ···················· | ···················· | ····················· | ···················· | ···················· | ···················· |
| A | 7052 | hCoV-19/India/UN-1111/2020\|EPI_ISL_421666\|2020-03- | ···················· | ···················· | ····················· | ···················· | ···················· | ···················· |
| A | 7053 | hCoV-19/India/UN-1115/2020\|EPI_ISL_421667\|2020-03- | ···················· | ···················· | ····················· | ···················· | ···················· | ···················· |
| A | 7054 | hCoV-19/India/UN-1125/2020\|EPI_ISL_421668\|2020-03- | ···················· | ···················· | ····················· | ···················· | ···················· | ···················· |
| A | 7055 | hCoV-19/India/UN-1616/2020\|EPI_ISL_421669\|2020-03- | ···················· | ···················· | ····················· | ···················· | ···················· | ···················· |
| A | 7056 | hCoV-19/India/UN-1617/2020\|EPI_ISL_421670\|2020-03- | ···················· | ···················· | ····················· | ···················· | ···················· | ···················· |
| A | 7057 | hCoV-19/India/UN-1621/2020\|EPI_ISL_421671\|2020-03- | ···················· | ···················· | ····················· | ···················· | ···················· | ···················· |
| A | 7058 | hCoV-19/India/UN-1644/2020\|EPI_ISL_421672\|2020-03- | ···················· | ···················· | ····················· | ···················· | ···················· | ···················· |
| A | 7059 | hCoV-19/USA/OH-OSUP0112/2020\|EPI_ISL_421675\|2020-0 | ···················· | ···················· | ····················· | ···················· | ···················· | ···················· |
| A | 7060 | hCoV-19/USA/MN124-MDH124/2020\|EPI_ISL_421683\|2020- | ···················· | ···················· | ····················· | ···················· | ···················· | ···················· |
| A | 7061 | hCoV-19/USA/MN128-MDH128/2020\|EPI_ISL_421684\|2020- | ···················· | ···················· | ····················· | ···················· | ···················· | ···················· |
| A | 7062 | hCoV-19/USA/MN129-MDH129/2020\|EPI_ISL_421685\|2020- | ···················· | ···················· | ····················· | ···················· | ···················· | ···················· |
| A | 7063 | hCoV-19/USA/MN130-MDH130/2020\|EPI_ISL_421686\|2020- | ···················· | ···················· | ····················· | ···················· | ···················· | ···················· |
| A | 7064 | hCoV-19/USA/MN131-MDH131/2020\|EPI_ISL_421687\|2020- | ···················· | ···················· | ····················· | ···················· | ···················· | ···················· |
| A | 7065 | hCoV-19/USA/MN132-MDH132/2020\|EPI_ISL_421688\|2020- | ···················· | ···················· | ····················· | ···················· | ···················· | ···················· |
| A | 7066 | hCoV-19/USA/MN133-MDH133/2020\|EPI_ISL_421689\|2020- | ···················· | ···················· | ····················· | ···················· | ···················· | ···················· |
| A | 7067 | hCoV-19/USA/MN134-MDH134/2020\|EPI_ISL_421690\|2020- | ···················· | ···················· | ····················· | ···················· | ···················· | ···················· |
| A | 7068 | hCoV-19/USA/MN135-MDH135/2020\|EPI_ISL_421691\|2020- | ···················· | ···················· | ····················· | ···················· | ···················· | ···················· |
| A | 7069 | hCoV-19/USA/MN136-MDH136/2020\|EPI_ISL_421692\|2020- | ···················· | ···················· | ····················· | ···················· | ···················· | ···················· |
| A | 7070 | hCoV-19/Netherlands/ZuidHolland_102/2020\|EPI_ISL_4 | ···················· | ···················· | ····················· | ···················· | ···················· | ···················· |
| A | 7071 | hCoV-19/Netherlands/ZuidHolland_103/2020\|EPI_ISL_4 | ···················· | ···················· | ····················· | ···················· | ···················· | ···················· |
| A | 7072 | hCoV-19/Netherlands/ZuidHolland_104/2020\|EPI_ISL_4 | ···················· | ···················· | ····················· | ···················· | ···················· | ···················· |
| A | 7073 | hCoV-19/Netherlands/ZuidHolland_32/2020\|EPI_ISL_42 | ···················· | ···················· | ····················· | ···················· | ···················· | ···················· |
| A | 7074 | hCoV-19/Netherlands/ZuidHolland_33/2020\|EPI_ISL_42 | ···················· | ···················· | ····················· | ···················· | ···················· | ···················· |
| A | 7075 | hCoV-19/Netherlands/ZuidHolland_35/2020\|EPI_ISL_42 | ···················· | ···················· | ····················· | ···················· | ···················· | ···················· |
| A | 7076 | hCoV-19/Netherlands/ZuidHolland_36/2020\|EPI_ISL_42 | ···················· | ···················· | ····················· | ···················· | ···················· | ···················· |
| A | 7077 | hCoV-19/Netherlands/ZuidHolland_37/2020\|EPI_ISL_42 | ···················· | ···················· | ····················· | ···················· | ···················· | ···················· |
| A | 7078 | hCoV-19/Netherlands/ZuidHolland_38/2020\|EPI_ISL_42 | ···················· | ···················· | ····················· | ···················· | ···················· | ···················· |
| A | 7079 | hCoV-19/Netherlands/ZuidHolland_39/2020\|EPI_ISL_42 | ···················· | ···················· | ····················· | ···················· | ···················· | ···················· |
| A | 7080 | hCoV-19/Netherlands/ZuidHolland_40/2020\|EPI_ISL_42 | ···················· | ···················· | ····················· | ···················· | ···················· | ···················· |
| A | 7081 | hCoV-19/Netherlands/ZuidHolland_41/2020\|EPI_ISL_42 | ···················· | ···················· | ····················· | ···················· | ···················· | ···················· |
| A | 7082 | hCoV-19/Netherlands/ZuidHolland_42/2020\|EPI_ISL_42 | ···················· | ···················· | ····················· | ···················· | ···················· | ···················· |
| A | 7083 | hCoV-19/Netherlands/ZuidHolland_43/2020\|EPI_ISL_42 | ···················· | ···················· | ····················· | ···················· | ···················· | ···················· |
| A | 7084 | hCoV-19/Netherlands/ZuidHolland_44/2020\|EPI_ISL_42 | ···················· | ···················· | ····················· | ···················· | ···················· | ···················· |
| A | 7085 | hCoV-19/Netherlands/ZuidHolland_45/2020\|EPI_ISL_42 | ···················· | ···················· | ····················· | ···················· | ···················· | ···················· |
| A | 7086 | hCoV-19/Netherlands/ZuidHolland_47/2020\|EPI_ISL_42 | ···················· | ···················· | ····················· | ···················· | ···················· | ···················· |
| A | 7087 | hCoV-19/Netherlands/ZuidHolland_48/2020\|EPI_ISL_42 | ········T··········· | ···················· | ····················· | ···················· | ···················· | ···················· |
| A | 7088 | hCoV-19/Netherlands/ZuidHolland_52/2020\|EPI_ISL_42 | ···················· | ···················· | ····················· | ···················· | ···················· | ···················· |
| A | 7089 | hCoV-19/Netherlands/ZuidHolland_55/2020\|EPI_ISL_42 | ···················· | ···················· | ····················· | ···················· | ···················· | ···················· |
| A | 7090 | hCoV-19/Netherlands/ZuidHolland_56/2020\|EPI_ISL_42 | ···················· | ···················· | ····················· | ···················· | ···················· | ···················· |
| A | 7091 | hCoV-19/Netherlands/ZuidHolland_57/2020\|EPI_ISL_42 | ···················· | ···················· | ····················· | ···················· | ···················· | ···················· |
| A | 7092 | hCoV-19/Netherlands/ZuidHolland_58/2020\|EPI_ISL_42 | ···················· | ···················· | ····················· | ···················· | ···················· | ···················· |
| A | 7093 | hCoV-19/Netherlands/ZuidHolland_61/2020\|EPI_ISL_42 | ···················· | ···················· | ····················· | ···················· | ···················· | ···················· |
| A | 7094 | hCoV-19/Netherlands/ZuidHolland_70/2020\|EPI_ISL_42 | ···················· | ···················· | ····················· | ···················· | ···················· | ···················· |
| A | 7095 | hCoV-19/Netherlands/ZuidHolland_72/2020\|EPI_ISL_42 | ···················· | ···················· | ····················· | ···················· | ···················· | ···················· |
| A | 7096 | hCoV-19/Netherlands/ZuidHolland_74/2020\|EPI_ISL_42 | ···················· | ···················· | ····················· | ···················· | ···················· | ···················· |
| A | 7097 | hCoV-19/Netherlands/ZuidHolland_75/2020\|EPI_ISL_42 | ···················· | ···················· | ····················· | ···················· | ···················· | ···················· |
| A | 7098 | hCoV-19/Netherlands/ZuidHolland_76/2020\|EPI_ISL_42 | ···················· | ···················· | ····················· | ···················· | ···················· | ···················· |
| A | 7099 | hCoV-19/Netherlands/ZuidHolland_77/2020\|EPI_ISL_42 | ···················· | ···················· | ····················· | ···················· | ···················· | ···················· |
| A | 7100 | hCoV-19/Netherlands/ZuidHolland_79/2020\|EPI_ISL_42 | ···················· | ···················· | ····················· | ···················· | ···················· | ···················· |
| A | 7101 | hCoV-19/Netherlands/ZuidHolland_80/2020\|EPI_ISL_42 | ···················· | ···················· | ····················· | ···················· | ···················· | ···················· |
| A | 7102 | hCoV-19/Netherlands/ZuidHolland_81/2020\|EPI_ISL_42 | ···················· | ···················· | ····················· | ···················· | ···················· | ···················· |
| A | 7103 | hCoV-19/Netherlands/ZuidHolland_82/2020\|EPI_ISL_42 | ···················· | ···················· | ····················· | ···················· | ···················· | ···················· |
| A | 7104 | hCoV-19/Netherlands/ZuidHolland_83/2020\|EPI_ISL_42 | ···················· | ···················· | ····················· | ···················· | ···················· | ···················· |
| A | 7105 | hCoV-19/USA/WA-UW-1279/2020\|EPI_ISL_422974\|2020-03 | ···················· | ···················· | ····················· | ···················· | ···················· | ···················· |
| A | 7106 | hCoV-19/USA/WA-UW-1327/2020\|EPI_ISL_422975\|2020-03 | ···················· | ···················· | ····················· | ···················· | ···················· | ···················· |
| A | 7107 | hCoV-19/USA/WA-UW-1306/2020\|EPI_ISL_422976\|2020-03 | ···················· | ···················· | ····················· | ···················· | ···················· | ···················· |

| | | | | | | | |
|---|---|---|---|---|---|---|---|
| A | 7108 hCoV-19/USA/WA-UW-1309/2020\|EPI_ISL_422977\|2020-03 | .................... | .................... | .................... | .................... | .................... | .................... |
| A | 7109 hCoV-19/USA/UN-UW-1291/2020\|EPI_ISL_422978\|2020-03 | .................... | .................... | .................... | .................... | .................... | .................... |
| A | 7110 hCoV-19/USA/UN-UW-1292/2020\|EPI_ISL_422979\|2020-03 | .................... | .................... | .................... | .................... | .................... | .................... |
| A | 7111 hCoV-19/USA/UN-UW-1307/2020\|EPI_ISL_422980\|2020-03 | .................... | .................... | .................... | .................... | .................... | .................... |
| A | 7112 hCoV-19/USA/WA-UW-1417/2020\|EPI_ISL_422981\|2020-03 | .................... | .................... | .................... | .................... | .................... | .................... |
| A | 7113 hCoV-19/USA/UN-UW-1403/2020\|EPI_ISL_422982\|2020-03 | .................... | .................... | .................... | .................... | .................... | .................... |
| A | 7114 hCoV-19/USA/UN-UW-1407/2020\|EPI_ISL_422983\|2020-03 | .................... | .................... | .................... | .................... | .................... | .................... |
| A | 7115 hCoV-19/USA/WA-UW-1300/2020\|EPI_ISL_422984\|2020-03 | .................... | .................... | .................... | .................... | .................... | .................... |
| A | 7116 hCoV-19/USA/WA-UW-1338/2020\|EPI_ISL_422986\|2020-03 | .................... | .................... | .................... | .................... | .................... | .................... |
| A | 7117 hCoV-19/USA/WA-UW-1353/2020\|EPI_ISL_422987\|2020-03 | .................... | .................... | .................... | .................... | .................... | .................... |
| A | 7118 hCoV-19/USA/WA-UW-1296/2020\|EPI_ISL_422988\|2020-03 | .................... | .................... | .................... | .................... | .................... | .................... |
| A | 7119 hCoV-19/USA/WA-UW-1297/2020\|EPI_ISL_422989\|2020-03 | .................... | .................... | .................... | .................... | .................... | .................... |
| A | 7120 hCoV-19/USA/WA-UW-1346/2020\|EPI_ISL_422996\|2020-03 | .................... | .................... | .................... | .................... | .................... | .................... |
| A | 7121 hCoV-19/USA/WA-UW-1351/2020\|EPI_ISL_422997\|2020-03 | .................... | .................... | .................... | .................... | .................... | .................... |
| A | 7122 hCoV-19/USA/WA-UW-1349/2020\|EPI_ISL_422999\|2020-03 | .................... | .................... | .................... | .................... | .................... | .................... |
| A | 7123 hCoV-19/USA/WI-UW-93/2020\|EPI_ISL_425150\|2020-04-0 | .................... | .................... | .................... | .................... | .................... | .................... |
| A | 7124 hCoV-19/USA/WI-UW-94/2020\|EPI_ISL_425151\|2020-03-3 | .................... | .................... | .................... | .................... | .................... | .................... |
| A | 7125 hCoV-19/USA/WI-UW-95/2020\|EPI_ISL_425152\|2020-03-3 | .................... | .................... | .................... | .................... | .................... | .................... |
| A | 7126 hCoV-19/USA/WI-UW-96/2020\|EPI_ISL_425153\|2020-04-0 | .................... | .................... | .................... | .................... | .................... | .................... |
| A | 7127 hCoV-19/USA/WI-UW-97/2020\|EPI_ISL_425154\|2020-03-3 | .................... | .................... | .................... | .................... | .................... | .................... |
| A | 7128 hCoV-19/USA/WI-UW-98/2020\|EPI_ISL_425155\|2020-03-3 | .................... | .................... | .................... | .................... | .................... | .................... |
| A | 7129 hCoV-19/USA/WI-UW-99/2020\|EPI_ISL_425156\|2020-04-0 | .................... | .................... | .................... | .................... | .................... | .................... |
| A | 7130 hCoV-19/USA/WI-UW-100/2020\|EPI_ISL_425157\|2020-04- | .................... | .................... | .................... | .................... | .................... | .................... |
| A | 7131 hCoV-19/USA/WI-UW-101/2020\|EPI_ISL_425158\|2020-03- | .................... | .................... | .................... | .................... | .................... | .................... |
| A | 7132 hCoV-19/USA/WI-UW-102/2020\|EPI_ISL_425159\|2020-03- | .................... | .................... | .................... | .................... | .................... | .................... |
| A | 7133 hCoV-19/USA/WI-UW-103/2020\|EPI_ISL_425160\|2020-04- | .................... | .................... | .................... | .................... | .................... | .................... |
| A | 7134 hCoV-19/USA/WI-UW-104/2020\|EPI_ISL_425161\|2020-04- | .................... | .................... | .................... | .................... | .................... | .................... |
| A | 7135 hCoV-19/USA/WI-UW-105/2020\|EPI_ISL_425162\|2020-03- | .................... | .................... | .................... | .................... | .................... | .................... |
| A | 7136 hCoV-19/USA/WI-UW-106/2020\|EPI_ISL_425163\|2020-03- | .................... | .................... | .................... | .................... | .................... | .................... |
| A | 7137 hCoV-19/USA/WI-UW-107/2020\|EPI_ISL_425164\|2020-03- | .................... | .................... | .................... | .................... | .................... | .................... |
| A | 7138 hCoV-19/USA/WI-UW-108/2020\|EPI_ISL_425165\|2020-03- | .................... | .................... | .................... | .................... | .................... | .................... |
| A | 7139 hCoV-19/USA/WI-UW-109/2020\|EPI_ISL_425166\|2020-03- | .................... | .................... | .................... | .................... | .................... | .................... |
| A | 7140 hCoV-19/USA/WI-UW-110/2020\|EPI_ISL_425167\|2020-03- | .................... | .................... | .................... | .................... | .................... | .................... |
| A | 7141 hCoV-19/USA/WI-UW-111/2020\|EPI_ISL_425168\|2020-03- | .................... | .................... | .................... | .................... | .................... | .................... |
| A | 7142 hCoV-19/USA/WI-UW-112/2020\|EPI_ISL_425169\|2020-03- | .................... | .................... | .................... | .................... | .................... | .................... |
| A | 7143 hCoV-19/USA/CO-UW-113/2020\|EPI_ISL_425170\|2020-03- | .................... | .................... | .................... | .................... | .................... | .................... |
| A | 7144 hCoV-19/USA/WI-UW-114/2020\|EPI_ISL_425171\|2020-03- | .................... | .................... | .................... | .................... | .................... | .................... |
| A | 7145 hCoV-19/USA/WI-UW-115/2020\|EPI_ISL_425172\|2020-03- | .................... | .................... | .................... | .................... | .................... | .................... |
| A | 7146 hCoV-19/USA/WI-UW-116/2020\|EPI_ISL_425173\|2020-03- | .................... | .................... | .................... | .................... | .................... | .................... |
| A | 7147 hCoV-19/USA/WI-UW-117/2020\|EPI_ISL_425174\|2020-03- | .................... | .................... | .................... | .................... | .................... | .................... |
| A | 7148 hCoV-19/USA/WI-UW-118/2020\|EPI_ISL_425175\|2020-03- | .................... | .................... | .................... | .................... | .................... | .................... |
| A | 7149 hCoV-19/USA/WI-UW-119/2020\|EPI_ISL_425176\|2020-03 | .................... | .................... | .................... | .................... | .................... | .................... |
| A | 7150 hCoV-19/Canada/ON_ON-VIDO-01-2/2020\|EPI_ISL_425177 | .................... | .A.................. | .................... | .................... | .................... | .................... |
| A | 7151 hCoV-19/Spain/VC-FISABIO-54/2020\|EPI_ISL_425178\|20 | .................... | .................... | .................... | .................... | .................... | .................... |
| A | 7152 hCoV-19/Spain/VC-FISABIO-55/2020\|EPI_ISL_425179\|20 | .................... | .................... | .................... | .................... | .................... | .................... |
| A | 7153 hCoV-19/Spain/VC-FISABIO-56/2020\|EPI_ISL_425180\|20 | .................... | .................... | .................... | .................... | .................... | .................... |
| A | 7154 hCoV-19/Spain/VC-FISABIO-57/2020\|EPI_ISL_425181\|20 | .................... | .................... | .................... | .................... | .................... | .................... |
| A | 7155 hCoV-19/Spain/VC-FISABIO-58/2020\|EPI_ISL_425182\|20 | .................... | .................... | .................... | .................... | .................... | .................... |
| A | 7156 hCoV-19/Spain/VC-FISABIO-59/2020\|EPI_ISL_425183\|20 | .................... | .................... | .................... | .................... | .................... | .................... |
| A | 7157 hCoV-19/Spain/VC-FISABIO-60/2020\|EPI_ISL_425184\|20 | .................... | .................... | .................... | .................... | .................... | .................... |
| A | 7158 hCoV-19/Spain/VC-FISABIO-61/2020\|EPI_ISL_425185\|20 | .................... | .................... | .................... | .................... | .................... | .................... |
| A | 7159 hCoV-19/Spain/VC-FISABIO-62/2020\|EPI_ISL_425186\|20 | .................... | .................... | .................... | .................... | .................... | .................... |
| A | 7160 hCoV-19/Spain/VC-FISABIO-63/2020\|EPI_ISL_425187\|20 | .................... | .................... | .................... | .................... | .................... | .................... |
| A | 7161 hCoV-19/Spain/VC-FISABIO-64/2020\|EPI_ISL_425188\|20 | .................... | .................... | .................... | .................... | .................... | .................... |
| A | 7162 hCoV-19/Spain/VC-FISABIO-65/2020\|EPI_ISL_425189\|20 | .................... | .................... | .................... | .................... | .................... | .................... |
| A | 7163 hCoV-19/Spain/VC-FISABIO-69/2020\|EPI_ISL_425193\|20 | .................... | .................... | .................... | .................... | .................... | .................... |
| A | 7164 hCoV-19/Spain/VC-FISABIO-72/2020\|EPI_ISL_425196\|20 | .................... | .................... | .................... | .................... | .................... | .................... |
| A | 7165 hCoV-19/Spain/VC-FISABIO-73/2020\|EPI_ISL_425197\|20 | .................... | .................... | .................... | .................... | .................... | .................... |
| A | 7166 hCoV-19/Spain/VC-FISABIO-75/2020\|EPI_ISL_425199\|20 | .................... | .................... | .................... | .................... | .................... | .................... |
| A | 7167 hCoV-19/USA/AZ-TG271872/2020\|EPI_ISL_426485\|2020-0 | .................... | .................... | .................... | .................... | .................... | .................... |
| A | 7168 hCoV-19/Germany/BY-ChVir-929-2/2020\|EPI_ISL_450198 | .................... | .................... | .................... | .................... | .................... | .................... |
| A | 7169 hCoV-19/Hangzhou/HZCDC0001/2020\|EPI_ISL_407313\|202 | .................... | .................... | .................... | .................... | .................... | .................... |
| A | 7170 hCoV-19/Netherlands/Gelderland_1/2020\|EPI_ISL_4144 | .................... | .................... | .................... | .................... | .................... | .................... |
| A | 7171 hCoV-19/Netherlands/Limburg_2/2020\|EPI_ISL_414424\| | .................... | .................... | .................... | .................... | .................... | .................... |
| A | 7172 hCoV-19/Netherlands/Limburg_3/2020\|EPI_ISL_414425\| | .................... | .................... | .................... | .................... | .................... | .................... |
| A | 7173 hCoV-19/Netherlands/Limburg_4/2020\|EPI_ISL_414426\| | .................... | .................... | .................... | .................... | .................... | .................... |
| A | 7174 hCoV-19/Netherlands/NoordBrabant_1/2020\|EPI_ISL_41 | .................... | .................... | .................... | .................... | .................... | .................... |
| A | 7175 hCoV-19/Netherlands/NoordBrabant_3/2020\|EPI_ISL_41 | .................... | .................... | .................... | .................... | .................... | .................... |
| A | 7176 hCoV-19/Netherlands/NoordBrabant_5/2020\|EPI_ISL_41 | .................... | .................... | .................... | .................... | .................... | .................... |
| A | 7177 hCoV-19/Netherlands/NoordBrabant_11/2020\|EPI_ISL_4 | .................... | .................... | .................... | .................... | .................... | .................... |
| A | 7178 hCoV-19/Netherlands/NoordHolland_1/2020\|EPI_ISL_41 | .................... | .................... | .................... | .................... | .................... | .................... |
| A | 7179 hCoV-19/Netherlands/Overijssel_1/2020\|EPI_ISL_4144 | .................... | .................... | .................... | .................... | .................... | .................... |
| A | 7180 hCoV-19/Netherlands/Utrecht_1/2020\|EPI_ISL_414435\| | .................... | .................... | .................... | .................... | .................... | .................... |
| A | 7181 hCoV-19/Netherlands/Utrecht_2/2020\|EPI_ISL_414436\| | .................... | .................... | .................... | .................... | .................... | .................... |
| A | 7182 hCoV-19/Netherlands/Utrecht_3/2020\|EPI_ISL_414437\| | .................... | .................... | .................... | .................... | .................... | .................... |
| A | 7183 hCoV-19/Netherlands/Utrecht_4/2020\|EPI_ISL_414438\| | .................... | .................... | .................... | .................... | .................... | .................... |
| A | 7184 hCoV-19/Netherlands/Utrecht_5/2020\|EPI_ISL_414439\| | .................... | .................... | .................... | .................... | .................... | .................... |
| A | 7185 hCoV-19/Netherlands/Utrecht_7/2020\|EPI_ISL_414440\| | .................... | .................... | .................... | .................... | .................... | .................... |

| | | | | | | | |
|---|---|---|---|---|---|---|---|
| A | 7186 hCoV-19/Netherlands/Utrecht_8/2020\|EPI_ISL_414441\| | ···················· | ···················· | ····················· | ···················· | ···················· | ···················· |
| A | 7187 hCoV-19/Netherlands/Utrecht_10/2020\|EPI_ISL_414442 | ···················· | ···················· | ····················· | ···················· | ···················· | ···················· |
| A | 7188 hCoV-19/Netherlands/Utrecht_11/2020\|EPI_ISL_414443 | ···················· | ···················· | ····················· | ···················· | ···················· | ···················· |
| A | 7189 hCoV-19/Netherlands/ZuidHolland_1/2020\|EPI_ISL_414 | ···················· | ···················· | ····················· | ···················· | ···················· | ···················· |
| A | 7190 hCoV-19/Netherlands/ZuidHolland_9/2020\|EPI_ISL_414 | ···················· | ···················· | ····················· | ···················· | ···················· | ···················· |
| A | 7191 hCoV-19/Netherlands/ZuidHolland_10/2020\|EPI_ISL_41 | ···················· | ···················· | ····················· | ···················· | ···················· | ···················· |
| A | 7192 hCoV-19/Netherlands/Limburg_5/2020\|EPI_ISL_414448\| | ···················· | ···················· | ····················· | ···················· | ···················· | ···················· |
| A | 7193 hCoV-19/Netherlands/NoordBrabant_2/2020\|EPI_ISL_41 | ···················· | ···················· | ····················· | ···················· | ···················· | ···················· |
| A | 7194 hCoV-19/Netherlands/NoordBrabant_6/2020\|EPI_ISL_41 | ···················· | ···················· | ····················· | ···················· | ···················· | ···················· |
| A | 7195 hCoV-19/Netherlands/NoordBrabant_12/2020\|EPI_ISL_4 | ···················· | ···················· | ····················· | ···················· | ···················· | ···················· |
| A | 7196 hCoV-19/Netherlands/NoordBrabant_14/2020\|EPI_ISL_4 | ···················· | ···················· | ····················· | ···················· | ···················· | ···················· |
| A | 7197 hCoV-19/Netherlands/NoordBrabant_16/2020\|EPI_ISL_4 | ···················· | ···················· | ····················· | ···················· | ···················· | ···················· |
| A | 7198 hCoV-19/Netherlands/NoordBrabant_17/2020\|EPI_ISL_4 | ···T················ | ···················· | ····················· | ···················· | ···················· | ···················· |
| A | 7199 hCoV-19/Netherlands/NoordBrabant_18/2020\|EPI_ISL_4 | ···················· | ···················· | ····················· | ···················· | ···················· | ···················· |
| A | 7200 hCoV-19/Netherlands/Overijssel_2/2020\|EPI_ISL_4144 | ···················· | ···················· | ····················· | ···················· | ···················· | ···················· |
| A | 7201 hCoV-19/Netherlands/Utrecht_6/2020\|EPI_ISL_414461\| | ···················· | ···················· | ····················· | ···················· | ···················· | ···················· |
| A | 7202 hCoV-19/Netherlands/Utrecht_12/2020\|EPI_ISL_414462 | ···················· | ···················· | ····················· | ···················· | ···················· | ···················· |
| A | 7203 hCoV-19/Netherlands/Utrecht_13/2020\|EPI_ISL_414463 | ···················· | ···················· | ····················· | ···················· | ···················· | ···················· |
| A | 7204 hCoV-19/Netherlands/Utrecht_14/2020\|EPI_ISL_414464 | ···················· | ···················· | ····················· | ···················· | ···················· | ···················· |
| A | 7205 hCoV-19/Netherlands/ZuidHolland_7/2020\|EPI_ISL_414 | ···················· | ···················· | ····················· | ···················· | ···················· | ···················· |
| A | 7206 hCoV-19/Netherlands/ZuidHolland_8/2020\|EPI_ISL_414 | ···················· | ···················· | ····················· | ···················· | ···················· | ···················· |
| A | 7207 hCoV-19/Netherlands/ZuidHolland_11/2020\|EPI_ISL_41 | ···················· | ···················· | ····················· | ···················· | ···················· | ···················· |
| A | 7208 hCoV-19/Netherlands/ZuidHolland_13/2020\|EPI_ISL_41 | ···················· | ···················· | ····················· | ···················· | ···················· | ···················· |
| A | 7209 hCoV-19/Netherlands/ZuidHolland_14/2020\|EPI_ISL_41 | ···················· | ···················· | ····················· | ···················· | ···················· | ···················· |
| A | 7210 hCoV-19/USA/NY1-PV08001/2020\|EPI_ISL_414476\|2020-0 | ···················· | ···················· | ····················· | ···················· | ···················· | ···················· |
| A | 7211 hCoV-19/Czech Republic/951/2020\|EPI_ISL_414477\|202 | ···················· | ···················· | ····················· | ···················· | ···················· | ···················· |
| A | 7212 hCoV-19/USA/CruiseA-19/2020\|EPI_ISL_414479\|2020-02 | ···················· | ···················· | ····················· | ···················· | ···················· | ···················· |
| A | 7213 hCoV-19/USA/CruiseA-21/2020\|EPI_ISL_414480\|2020-02 | ···················· | ···················· | ····················· | ···················· | ···················· | ···················· |
| A | 7214 hCoV-19/USA/CruiseA-22/2020\|EPI_ISL_414481\|2020-02 | ···················· | ···················· | ····················· | ···················· | ···················· | ···················· |
| A | 7215 hCoV-19/USA/CruiseA-23/2020\|EPI_ISL_414482\|2020-02 | ···················· | ···················· | ····················· | ···················· | ···················· | ···················· |
| A | 7216 hCoV-19/USA/CruiseA-24/2020\|EPI_ISL_414483\|2020-02 | ···················· | ···················· | ····················· | ···················· | ···················· | ···················· |
| A | 7217 hCoV-19/USA/CruiseA-25/2020\|EPI_ISL_414484\|2020-02 | ···················· | ···················· | ····················· | ···················· | ···················· | ···················· |
| A | 7218 hCoV-19/USA/CruiseA-26/2020\|EPI_ISL_414485\|2020-02 | ···················· | ···················· | ····················· | ···················· | ···················· | ···················· |
| A | 7219 hCoV-19/Ireland/CO-NVRL-20134/2020\|EPI_ISL_414487\| | ···················· | ···················· | ····················· | ···················· | ···················· | ···················· |
| A | 7222 hCoV-19/Germany/NW-HHU-02-1/2020\|EPI_ISL_414497\|20 | ···················· | ···················· | ····················· | ···················· | ···················· | ···················· |
| A | 7223 hCoV-19/Germany/NW-HHU-03/2020\|EPI_ISL_414498\|2020 | ···················· | ···················· | ····················· | ···················· | ···················· | ···················· |
| A | 7224 hCoV-19/Germany/NW-HHU-04/2020\|EPI_ISL_414499\|2020 | ···················· | ···················· | ····················· | ···················· | ···················· | ···················· |
| A | 7225 hCoV-19/Peru/LIM-010/2020\|EPI_ISL_415787\|2020-03-1 | ···················· | ···················· | ····················· | ···················· | ···················· | ···················· |
| A | 7226 hCoV-19/England/SHEF-C0190/2020\|EPI_ISL_420257\|202 | ···················· | ···················· | ····················· | ···················· | ···················· | ···················· |
| A | 7228 hCoV-19/England/SHEF-C0181/2020\|EPI_ISL_420260\|202 | ···················· | ···················· | ····················· | ···················· | ···················· | ···················· |
| A | 7229 hCoV-19/England/SHEF-C0725/2020\|EPI_ISL_420261\|202 | ···················· | ···················· | ····················· | ···················· | ···················· | ···················· |
| A | 7230 hCoV-19/England/SHEF-C0752/2020\|EPI_ISL_420262\|202 | ···················· | ···················· | ····················· | ···················· | ···················· | ···················· |
| A | 7231 hCoV-19/England/SHEF-C04E2/2020\|EPI_ISL_420263\|202 | ···················· | ···················· | ····················· | ···················· | ···················· | ···················· |
| A | 7232 hCoV-19/England/SHEF-C06FB/2020\|EPI_ISL_420264\|202 | ···················· | ···················· | ····················· | ···················· | ···················· | ···················· |
| A | 7233 hCoV-19/England/SHEF-C078F/2020\|EPI_ISL_420265\|202 | ···················· | ···················· | ····················· | ···················· | ···················· | ···················· |
| A | 7234 hCoV-19/England/SHEF-C0057/2020\|EPI_ISL_420266\|202 | ···················· | ···················· | ····················· | ···················· | ···················· | ···················· |
| A | 7235 hCoV-19/England/SHEF-C00DF/2020\|EPI_ISL_420267\|202 | ···················· | ···················· | ····················· | ···················· | ···················· | ···················· |
| A | 7236 hCoV-19/England/SHEF-C0093/2020\|EPI_ISL_420268\|202 | ···················· | ···················· | ····················· | ···················· | ···················· | ···················· |
| A | 7237 hCoV-19/England/SHEF-C04F1/2020\|EPI_ISL_420269\|202 | ···················· | ···················· | ····················· | ···················· | ···················· | ···················· |
| A | 7238 hCoV-19/England/SHEF-C0707/2020\|EPI_ISL_420270\|202 | ···················· | ···················· | ····················· | ···················· | ···················· | ···················· |
| A | 7239 hCoV-19/England/SHEF-C0075/2020\|EPI_ISL_420271\|202 | ···················· | ···················· | ····················· | ···················· | ···················· | ···················· |
| A | 7240 hCoV-19/England/SHEF-C002A/2020\|EPI_ISL_420272\|202 | ···················· | ···················· | ····················· | ···················· | ···················· | ···················· |
| A | 7241 hCoV-19/England/SHEF-C000C/2020\|EPI_ISL_420273\|202 | ···················· | ···················· | ····················· | ···················· | ···················· | ···················· |
| A | 7242 hCoV-19/England/SHEF-C00EE/2020\|EPI_ISL_420274\|202 | ···················· | ···················· | ····················· | ···················· | ···················· | ···················· |
| A | 7243 hCoV-19/England/SHEF-C0039/2020\|EPI_ISL_420275\|202 | ···················· | ···················· | ····················· | ···················· | ···················· | ···················· |
| A | 7244 hCoV-19/England/SHEF-C00B1/2020\|EPI_ISL_420276\|202 | ···················· | ···················· | ····················· | ···················· | ···················· | ···················· |
| A | 7245 hCoV-19/England/SHEF-C0066/2020\|EPI_ISL_420277\|202 | ···················· | ···················· | ····················· | ···················· | ···················· | ···················· |
| A | 7246 hCoV-19/England/SHEF-C0646/2020\|EPI_ISL_420278\|202 | ···················· | ···················· | ····················· | ···················· | ···················· | ···················· |
| A | 7247 hCoV-19/USA/NY-NYUMC89/2020\|EPI_ISL_421590\|2020-03 | ···················· | ···················· | ····················· | ···················· | ···················· | ···················· |
| A | 7248 hCoV-19/USA/NY-NYUMC90/2020\|EPI_ISL_421591\|2020-03 | ···················· | ···················· | ····················· | ···················· | ···················· | ···················· |
| A | 7249 hCoV-19/USA/NY-NYUMC91/2020\|EPI_ISL_421592\|2020-03 | ···················· | ···················· | ····················· | ···················· | ···················· | ···················· |
| A | 7250 hCoV-19/USA/NY-PV09006/2020\|EPI_ISL_421593\|2020-03 | ···················· | ···················· | ····················· | ···················· | ···················· | ···················· |
| A | 7251 hCoV-19/USA/NY-PV09019/2020\|EPI_ISL_421594\|2020-03 | ···················· | ···················· | ····················· | ···················· | ···················· | ···················· |
| A | 7252 hCoV-19/USA/NY-PV09020/2020\|EPI_ISL_421595\|2020-03 | ···················· | ···················· | ····················· | ···················· | ···················· | ···················· |
| A | 7253 hCoV-19/USA/NY-PV09021/2020\|EPI_ISL_421596\|2020-03 | ···················· | ···················· | ····················· | ···················· | ···················· | ···················· |
| A | 7254 hCoV-19/USA/NY-PV09023/2020\|EPI_ISL_421597\|2020-03 | ···················· | ···················· | ····················· | ···················· | ···················· | ···················· |
| A | 7255 hCoV-19/USA/NY-PV09025/2020\|EPI_ISL_421598\|2020-03 | ···················· | ···················· | ····················· | ···················· | ···················· | ···················· |
| A | 7256 hCoV-19/USA/NY-PV09028/2020\|EPI_ISL_421599\|2020-03 | ···················· | ···················· | ····················· | ···················· | ···················· | ···················· |
| A | 7257 hCoV-19/USA/NY-PV09029/2020\|EPI_ISL_421600\|2020-03 | ···················· | ···················· | ····················· | ···················· | ···················· | ···················· |
| A | 7258 hCoV-19/USA/NY-PV09032/2020\|EPI_ISL_421602\|2020-03 | ···················· | ···················· | ····················· | ···················· | ···················· | ···················· |
| A | 7259 hCoV-19/USA/NY-PV09037/2020\|EPI_ISL_421603\|2020-03 | ···················· | ···················· | ····················· | ···················· | ···················· | ···················· |
| A | 7260 hCoV-19/USA/NY-PV09039/2020\|EPI_ISL_421604\|2020-03 | ···················· | ···················· | ····················· | ···················· | ···················· | ···················· |
| A | 7261 hCoV-19/USA/NY-PV09041/2020\|EPI_ISL_421605\|2020-03 | ···················· | ···················· | ····················· | ···················· | ···················· | ···················· |
| A | 7262 hCoV-19/USA/NY-PV09043/2020\|EPI_ISL_421606\|2020-03 | ···················· | ···················· | ····················· | ···················· | ···················· | ···················· |
| A | 7263 hCoV-19/USA/NY-PV09046/2020\|EPI_ISL_421608\|2020-03 | ···················· | ···················· | ····················· | ···················· | ···················· | ···················· |
| A | 7264 hCoV-19/Netherlands/ZuidHolland_62/2020\|EPI_ISL_42 | ···················· | ···················· | ····················· | ···················· | ···················· | ···················· |
| A | 7265 hCoV-19/Netherlands/ZuidHolland_64/2020\|EPI_ISL_42 | ···················· | ···················· | ····················· | ···················· | ···················· | ···················· |
| A | 7266 hCoV-19/Netherlands/ZuidHolland_66/2020\|EPI_ISL_42 | ···················· | ···················· | ····················· | ···················· | ···················· | ···················· |

A 7267 hCoV-19/Netherlands/ZuidHolland_67/2020|EPI_ISL_42 ···················· ···················· ···················· ···················· ···················· ····················
A 7268 hCoV-19/Netherlands/ZuidHolland_71/2020|EPI_ISL_42 ···················· ···················· ···················· ···················· ···················· ····················
A 7269 hCoV-19/South Korea/KCDC2001/2020|EPI_ISL_425117|2 ···················· ···················· ···················· ···················· ···················· ····················
A 7270 hCoV-19/South Korea/KCDC2002/2020|EPI_ISL_425118|2 ···················· ···················· ···················· ···················· ···················· ····················
A 7271 hCoV-19/Germany/NW-HHU-37/2020|EPI_ISL_425121|2020 ···················· ···················· ···················· ···················· ···················· ····················
A 7272 hCoV-19/Germany/NW-HHU-38/2020|EPI_ISL_425122|2020 ···················· ···················· ···················· ···················· ···················· ····················
A 7273 hCoV-19/Germany/NW-HHU-39/2020|EPI_ISL_425123|2020 ···················· ···················· ···················· ···················· ···················· G···················
A 7274 hCoV-19/Germany/NW-HHU-40/2020|EPI_ISL_425124|2020 ···················· ···················· ···················· ···················· ···················· ····················
A 7275 hCoV-19/Germany/NW-HHU-41/2020|EPI_ISL_425125|2020 ···················· ···················· ···················· ···················· ···················· ····················
A 7276 hCoV-19/Germany/NW-HHU-42/2020|EPI_ISL_425126|2020 ···················· ···················· ···················· ···················· ···················· ····················
A 7277 hCoV-19/Germany/NW-HHU-42-2/2020|EPI_ISL_425127|20 ···················· ···················· ···················· ···················· ···················· ····················
A 7278 hCoV-19/Germany/NW-HHU-42-3/2020|EPI_ISL_425128|20 ···················· ···················· ···················· ···················· ··············N····· ····················
A 7279 hCoV-19/Germany/NW-HHU-46/2020|EPI_ISL_425130|2020 ···················· ···················· ···················· ···················· ···················· ····················
A 7280 hCoV-19/Germany/NW-HHU-47/2020|EPI_ISL_425131|2020 ···················· ···················· ···················· ···················· ···················· ····················
A 7281 hCoV-19/Germany/NW-HHU-48/2020|EPI_ISL_425132|2020 ···················· ···················· ···················· ···················· ···················· ····················
A 7282 hCoV-19/Germany/NW-HHU-43/2020|EPI_ISL_425138|2020 ···················· ···················· ···················· ···················· ···················· ····················
A 7283 hCoV-19/Germany/NW-HHU-44/2020|EPI_ISL_425139|2020 ···················· ···················· ···················· ···················· ···················· ····················
A 7284 hCoV-19/USA/WI-UW-85/2020|EPI_ISL_425142|2020-04-0 ···················· ···················· ···················· ···················· ···················· ····················
A 7285 hCoV-19/USA/WI-UW-86/2020|EPI_ISL_425143|2020-04-0 ···················· ···················· ···················· ···················· ···················· ····················
A 7286 hCoV-19/USA/WI-UW-87/2020|EPI_ISL_425144|2020-04-0 ···················· ···················· ···················· ···················· ···················· ····················
A 7287 hCoV-19/USA/WI-UW-88/2020|EPI_ISL_425145|2020-04-0 ···················· ···················· ···················· ···················· ···················· ····················
A 7288 hCoV-19/USA/WI-UW-89/2020|EPI_ISL_425146|2020-03-3 ···················· ···················· ···················· ···················· ···················· ····················
A 7289 hCoV-19/USA/WI-UW-90/2020|EPI_ISL_425147|2020-04-0 ···················· ···················· ···················· ···················· ···················· ····················
A 7290 hCoV-19/USA/WI-UW-91/2020|EPI_ISL_425148|2020-04-0 ···················· ···················· ···················· ···················· ···················· ····················
A 7291 hCoV-19/USA/WI-UW-92/2020|EPI_ISL_425149|2020-03-3 ···················· ···················· ···················· ···················· ···················· ····················
A 7292 hCoV-19/India/GJ-GBRC1/2020|EPI_ISL_426414|2020-04 ···················· ···················· ···················· ···················· ···················· ····················
A 7293 hCoV-19/India/GJ-GBRC1s/2020|EPI_ISL_426415|2020-0 ···················· ···················· ···················· ···················· ···················· ····················
A 7294 hCoV-19/USA/CT_9849/2020|EPI_ISL_426416|2020-03-08 ···················· ···················· ···················· ···················· ···················· ····················
A 7295 hCoV-19/USA/GA_2070/2020|EPI_ISL_426417|2020-03-07 ···················· ···················· ···················· ···················· ···················· ····················
A 7296 hCoV-19/USA/GA_2147/2020|EPI_ISL_426418|2020-03 ···················· ···················· ···················· ···················· ···················· ····················
A 7297 hCoV-19/USA/GA_2275/2020|EPI_ISL_426419|2020-03 ···················· ···················· ···················· ···················· ···················· ····················
A 7298 hCoV-19/USA/HI_4970/2020|EPI_ISL_426420|2020-03-05 ···················· ·A·················· ···················· ···················· ···················· ····················
A 7299 hCoV-19/USA/HI_7881/2020|EPI_ISL_426421|2020-03-07 ···················· ···················· ···················· ···················· ···················· ····················
A 7300 hCoV-19/USA/MD_0027/2020|EPI_ISL_426425|2020-03-04 ···················· ···················· ···················· ···················· ···················· ····················
A 7301 hCoV-19/USA/MN_0100/2020|EPI_ISL_426426|2020-03-05 ···················· ···················· ···················· ···················· ···················· ····················
A 7302 hCoV-19/USA/MN_0101/2020|EPI_ISL_426427|2020-03-07 ···················· ···················· ···················· ···················· ···················· ····················
A 7303 hCoV-19/USA/NC_0025/2020|EPI_ISL_426428|2020-03-08 ···················· ···················· ···················· ···················· ···················· ····················
A 7304 hCoV-19/USA/NV_0016/2020|EPI_ISL_426429|2020-03-07 ···················· ···················· ···················· ···················· ···················· ····················
A 7305 hCoV-19/USA/OH_0019/2020|EPI_ISL_426430|2020-03-08 ···················· ···················· ···················· ···················· ···················· ····················
A 7306 hCoV-19/USA/OH_0020/2020|EPI_ISL_426431|2020-03-07 ···················· ···················· ···················· ···················· ···················· ····················
A 7307 hCoV-19/USA/PA_2734/2020|EPI_ISL_426432|2020-03-07 ···················· ···················· ···················· ···················· ···················· ····················
A 7308 hCoV-19/USA/PA_2735/2020|EPI_ISL_426433|2020-03-07 ···················· ···················· ···················· ···················· ···················· ····················
A 7309 hCoV-19/USA/PA_2887/2020|EPI_ISL_426434|2020-03-08 ···················· ···················· ···················· ···················· ···················· ····················
A 7310 hCoV-19/USA/RI_0882/2020|EPI_ISL_426435|2020-03-09 ···················· ···················· ···················· ···················· ···················· ····················
A 7311 hCoV-19/USA/WA_0418/2020|EPI_ISL_426436|2020-03-19 ···················· ···················· ···················· ···················· ···················· ····················
A 7312 hCoV-19/USA/WA_0422/2020|EPI_ISL_426437|2020-03-19 ···················· ···················· ···················· ···················· ···················· ····················
A 7313 hCoV-19/USA/WA_0427/2020|EPI_ISL_426438|2020-03-19 ···················· ···················· ···················· ···················· ···················· ····················
A 7314 hCoV-19/USA/WA_0428/2020|EPI_ISL_426439|2020-03-19 ···················· ···················· ···················· ···················· ···················· ····················
A 7315 hCoV-19/USA/WA_0429/2020|EPI_ISL_426440|2020-03-19 ···················· ···················· ···················· ···················· ···················· ····················
A 7316 hCoV-19/USA/WA_0432/2020|EPI_ISL_426441|2020-03-19 ···················· ···················· ···················· ···················· ···················· ····················
A 7317 hCoV-19/USA/WA_0434/2020|EPI_ISL_426442|2020-03-19 ···················· ···················· ···················· ···················· ···················· ····················
A 7318 hCoV-19/USA/WA_0440/2020|EPI_ISL_426443|2020-03-19 ···················· ···················· ···················· ···················· ···················· ····················
A 7319 hCoV-19/USA/WA_0441/2020|EPI_ISL_426444|2020-03-19 ···················· ···················· ···················· ···················· ···················· ····················
A 7320 hCoV-19/USA/WA_0442/2020|EPI_ISL_426445|2020-03-19 ···················· ···················· ···················· ···················· ···················· ····················
A 7321 hCoV-19/USA/WA_0447/2020|EPI_ISL_426446|2020-03-19 ···················· ···················· ···················· ···················· ···················· ····················
A 7322 hCoV-19/USA/WA_0448/2020|EPI_ISL_426447|2020-03-19 ···················· ···················· ···················· ···················· ···················· ····················
A 7323 hCoV-19/USA/WA_0457/2020|EPI_ISL_426448|2020-03-20 ···················· ···················· ···················· ···················· ···················· ····················
A 7324 hCoV-19/USA/WA_0478/2020|EPI_ISL_426449|2020-03-19 ···················· ···················· ···················· ···················· ···················· ····················
A 7325 hCoV-19/USA/WA_0480/2020|EPI_ISL_426450|2020-03-19 ···················· ···················· ···················· ···················· ···················· ····················
A 7326 hCoV-19/USA/WA_0484/2020|EPI_ISL_426451|2020-03-19 ···················· ···················· ···················· ···················· ···················· ····················
A 7327 hCoV-19/USA/WA_0711/2020|EPI_ISL_426452|2020-03-19 ···················· ···················· ···················· ···················· ···················· ····················
A 7328 hCoV-19/USA/WA_0797/2020|EPI_ISL_426453|2020-03-20 ···················· ···················· ···················· ···················· ···················· ····················
A 7329 hCoV-19/USA/VA-DCLS-0041/2020|EPI_ISL_426454|2020- ···················· ···················· ···················· ···················· ···················· ····················
A 7330 hCoV-19/USA/VA-DCLS-0047/2020|EPI_ISL_426455|2020- ···················· ···················· ···················· ···················· ···················· ····················
A 7331 hCoV-19/USA/VA-DCLS-0048/2020|EPI_ISL_426456|2020- ···················· ···················· ···················· ···················· ···················· ····················
A 7332 hCoV-19/USA/VA-DCLS-0049/2020|EPI_ISL_426457|2020- ···················· ···················· ···················· ···················· ···················· ····················
A 7333 hCoV-19/USA/VA-DCLS-0050/2020|EPI_ISL_426458|2020- ···················· ···················· ···················· ···················· ···················· ····················
A 7334 hCoV-19/USA/VA-DCLS-0051/2020|EPI_ISL_426459|2020- ···················· ···················· ···················· ···················· ···················· ····················
A 7335 hCoV-19/USA/VA-DCLS-0052/2020|EPI_ISL_426460|2020- ···················· ···················· ···················· ···················· ···················· ····················
A 7336 hCoV-19/USA/VA-DCLS-0053/2020|EPI_ISL_426461|2020- ···················· ···················· ···················· ···················· ···················· ····················
A 7337 hCoV-19/USA/VA-DCLS-0054/2020|EPI_ISL_426462|2020- ···················· ···················· ···················· ···················· ···················· ····················
A 7338 hCoV-19/USA/VA-DCLS-0055/2020|EPI_ISL_426463|2020- ···················· ···················· ···················· ···················· ···················· ····················
A 7339 hCoV-19/USA/VA-DCLS-0056/2020|EPI_ISL_426464|2020- ···················· ···················· ···················· ··············A····· ···················· ····················
A 7340 hCoV-19/USA/VA-DCLS-0057/2020|EPI_ISL_426465|2020- ···················· ···················· ···················· ···················· ···················· ····················
A 7341 hCoV-19/USA/VA-DCLS-0058/2020|EPI_ISL_426466|2020- ···················· ···················· ···················· ···················· ···················· ····················
A 7342 hCoV-19/USA/VA-DCLS-0059/2020|EPI_ISL_426467|2020- ···················· ···················· ···················· ···················· ···················· ····················
A 7343 hCoV-19/USA/VA-DCLS-0060/2020|EPI_ISL_426468|2020- ···················· ···················· ···················· ···················· ···················· ····················
A 7344 hCoV-19/USA/VA-DCLS-0061/2020|EPI_ISL_426469|2020- ···················· ···················· ···················· ···················· ···················· ····················

| | | | | | | | | | |
|---|---|---|---|---|---|---|---|---|---|
| A | 7345 | hCoV-19/USA/VA-DCLS-0063/2020\|EPI_ISL_426470\|2020- | .................... | .................... | .................... | .................... | .................... | .................... | |
| A | 7346 | hCoV-19/USA/VA-DCLS-0064/2020\|EPI_ISL_426471\|2020- | .................... | .................... | .................... | .................... | .................... | .................... | |
| A | 7347 | hCoV-19/USA/VA-DCLS-0065/2020\|EPI_ISL_426472\|2020- | .................... | .................... | .................... | .................... | .................... | .................... | |
| A | 7348 | hCoV-19/USA/VA-DCLS-0066/2020\|EPI_ISL_426473\|2020- | .................... | .................... | .................... | .................... | .................... | .................... | |
| A | 7349 | hCoV-19/USA/VA-DCLS-0067/2020\|EPI_ISL_426474\|2020- | .................... | .................... | .................... | .................... | .................... | .................... | |
| A | 7350 | hCoV-19/USA/VA-DCLS-0068/2020\|EPI_ISL_426475\|2020- | .................... | .................... | .................... | .................... | .................... | .................... | |
| A | 7351 | hCoV-19/Australia/NSW213/2020\|EPI_ISL_427726\|2020- | .................... | .................... | .................... | .................... | .................... | .................... | |
| A | 7352 | hCoV-19/Australia/NSW214/2020\|EPI_ISL_427727\|2020- | .................... | .................... | .................... | .................... | .................... | .................... | |
| A | 7353 | hCoV-19/Australia/NSW113/2020\|EPI_ISL_427728\|2020- | .................... | .................... | .................... | .................... | .................... | .................... | |
| A | 7354 | hCoV-19/Australia/NSW77/2020\|EPI_ISL_427729\|2020-0 | .................... | .................... | .................... | .................... | .................... | .................... | |
| A | 7355 | hCoV-19/Australia/NSW215/2020\|EPI_ISL_427730\|2020- | .................... | .................... | .................... | .................... | .................... | .................... | |
| A | 7356 | hCoV-19/Australia/NSW165/2020\|EPI_ISL_427731\|2020- | .................... | .................... | .................... | .................... | .................... | .................... | |
| A | 7357 | hCoV-19/Australia/NSW216/2020\|EPI_ISL_427732\|2020- | .................... | .................... | .................... | .................... | .................... | .................... | |
| A | 7358 | hCoV-19/Australia/NSW67/2020\|EPI_ISL_427733\|2020-0 | .................... | .................... | .................... | .................... | .................... | .................... | |
| A | 7359 | hCoV-19/Australia/NSW176/2020\|EPI_ISL_427734\|2020- | .................... | .................... | .................... | .................... | .................... | .................... | |
| A | 7360 | hCoV-19/Australia/NSW158/2020\|EPI_ISL_427735\|2020- | .................... | .................... | .................... | .................... | .................... | .................... | |
| A | 7361 | hCoV-19/Australia/NSW190/2020\|EPI_ISL_427736\|2020- | .................... | .................... | .................... | .................... | .................... | .................... | |
| A | 7362 | hCoV-19/Australia/NSW143/2020\|EPI_ISL_427737\|2020- | .................... | .................... | .................... | .................... | .................... | .................... | |
| A | 7363 | hCoV-19/Australia/NSW144/2020\|EPI_ISL_427738\|2020- | .................... | .................... | .................... | .................... | .................... | .................... | |
| A | 7364 | hCoV-19/Australia/NSW168/2020\|EPI_ISL_427739\|2020- | .................... | .................... | .................... | .................... | .................... | .................... | |
| A | 7365 | hCoV-19/Australia/NSW189/2020\|EPI_ISL_427740\|2020- | .................... | .................... | .................... | .................... | .................... | .................... | |
| A | 7366 | hCoV-19/Australia/NSW110/2020\|EPI_ISL_427741\|2020- | .................... | .................... | .................... | .................... | .................... | .................... | |
| A | 7367 | hCoV-19/Australia/NSW134/2020\|EPI_ISL_427742\|2020- | .................... | .................... | .................... | .................... | .................... | .................... | |
| A | 7368 | hCoV-19/Australia/NSW171/2020\|EPI_ISL_427743\|2020- | .................... | .................... | .................... | .................... | .................... | .................... | |
| A | 7369 | hCoV-19/Australia/NSW72/2020\|EPI_ISL_427744\|2020-0 | .................... | .................... | .................... | .................... | .................... | .................... | |
| A | 7370 | hCoV-19/Australia/NSW205/2020\|EPI_ISL_427745\|2020- | .................... | .................... | .................... | .................... | .................... | .................... | |
| A | 7371 | hCoV-19/Australia/NSW197/2020\|EPI_ISL_427746\|2020- | .................... | .................... | .................... | .................... | .................... | .................... | |
| A | 7372 | hCoV-19/Australia/NSW199/2020\|EPI_ISL_427747\|2020- | .................... | .................... | .................... | .................... | .................... | .................... | |
| A | 7373 | hCoV-19/Australia/NSW106/2020\|EPI_ISL_427748\|2020- | .................... | .................... | .................... | .................... | .................... | .................... | |
| A | 7374 | hCoV-19/Australia/NSW142/2020\|EPI_ISL_427749\|2020- | .................... | .................... | .................... | .................... | .................... | .................... | |
| A | 7375 | hCoV-19/Australia/NSW73/2020\|EPI_ISL_427750\|2020-0 | .................... | .................... | .................... | .................... | .................... | .................... | |
| A | 7376 | hCoV-19/Australia/NSW68/2020\|EPI_ISL_427751\|2020-0 | .................... | .................... | .................... | .................... | .................... | .................... | |
| A | 7377 | hCoV-19/Australia/NSW184/2020\|EPI_ISL_427752\|2020- | .................... | .................... | .................... | .................... | .................... | .................... | |
| A | 7378 | hCoV-19/Australia/NSW208/2020\|EPI_ISL_427753\|2020- | .................... | .................... | .................... | .................... | .................... | .................... | |
| A | 7379 | hCoV-19/Australia/NSW109/2020\|EPI_ISL_427754\|2020- | .................... | .................... | .................... | .................... | .................... | .................... | |
| A | 7380 | hCoV-19/Australia/NSW209/2020\|EPI_ISL_427755\|2020- | .................... | .................... | .................... | .................... | .................... | .................... | |
| A | 7381 | hCoV-19/Australia/NSW117/2020\|EPI_ISL_427756\|2020- | .................... | .................... | .................... | .................... | .................... | .................... | |
| A | 7382 | hCoV-19/Australia/NSW70/2020\|EPI_ISL_427757\|2020-0 | .................... | .................... | .................... | .................... | .................... | .................... | |
| A | 7383 | hCoV-19/Australia/NSW162/2020\|EPI_ISL_427758\|2020- | .................... | .................... | .................... | .................... | .................... | .................... | |
| A | 7384 | hCoV-19/Australia/NSW71/2020\|EPI_ISL_427759\|2020-0 | .................... | .................... | .................... | .................... | .................... | .................... | |
| A | 7385 | hCoV-19/Australia/NSW129/2020\|EPI_ISL_427760\|2020- | .................... | .................... | .................... | .................... | .................... | .................... | |
| A | 7386 | hCoV-19/Australia/NSW138/2020\|EPI_ISL_427761\|2020- | .................... | .................... | .................... | .................... | .................... | .................... | |
| A | 7387 | hCoV-19/Australia/NSW202/2020\|EPI_ISL_427762\|2020- | .................... | .................... | .................... | .................... | .........T.......... | .................... | |
| A | 7388 | hCoV-19/Australia/NSW194/2020\|EPI_ISL_427763\|2020- | .................... | .................... | .................... | .................... | .................... | .................... | |
| A | 7389 | hCoV-19/Australia/NSW147/2020\|EPI_ISL_427764\|2020- | .................... | .................... | .................... | .................... | .................... | .................... | |
| A | 7390 | hCoV-19/Australia/NSW203/2020\|EPI_ISL_427765\|2020- | .................... | .................... | .................... | .................... | .................... | .................... | |
| A | 7391 | hCoV-19/Australia/NSW210/2020\|EPI_ISL_427766\|2020- | .................... | .................... | .................... | .................... | .................... | .................... | |
| A | 7392 | hCoV-19/Australia/NSW195/2020\|EPI_ISL_427767\|2020- | .................... | .................... | .................... | .................... | .................... | .................... | |
| A | 7393 | hCoV-19/Australia/NSW130/2020\|EPI_ISL_427768\|2020- | .................... | .................... | .................... | .................... | .................... | .................... | |
| A | 7394 | hCoV-19/Australia/NSW97/2020\|EPI_ISL_427769\|2020-0 | .................... | .................... | .................... | .................... | .................... | .................... | |
| A | 7395 | hCoV-19/Australia/NSW78/2020\|EPI_ISL_427770\|2020-0 | .................... | .................... | .................... | .................... | .................... | .................... | |
| A | 7396 | hCoV-19/Australia/NSW166/2020\|EPI_ISL_427771\|2020- | .................... | .................... | .................... | .................... | .................... | .................... | |
| A | 7397 | hCoV-19/Australia/NSW107/2020\|EPI_ISL_427772\|2020- | .................... | .................... | .................... | .................... | .................... | .................... | |
| A | 7398 | hCoV-19/Australia/NSW64/2020\|EPI_ISL_427773\|2020-0 | .................... | .................... | .................... | .................... | .................... | .................... | |
| A | 7399 | hCoV-19/Australia/NSW98/2020\|EPI_ISL_427774\|2020-0 | .................... | .................... | .................... | .................... | .................... | .................... | |
| A | 7400 | hCoV-19/Australia/NSW149/2020\|EPI_ISL_427775\|2020- | .................... | .................... | .................... | .................... | .................... | .................... | |
| A | 7401 | hCoV-19/Australia/NSW89/2020\|EPI_ISL_427776\|2020-0 | .................... | .................... | .................... | .................... | .................... | .................... | |
| A | 7402 | hCoV-19/Australia/NSW99/2020\|EPI_ISL_427777\|2020-0 | .................... | .................... | .................... | .................... | .................... | .................... | |
| A | 7403 | hCoV-19/Australia/NSW122/2020\|EPI_ISL_427778\|2020- | .................... | .................... | .................... | .................... | .................... | .................... | |
| A | 7404 | hCoV-19/Australia/NSW169/2020\|EPI_ISL_427779\|2020- | .................... | .................... | .................... | .................... | .................... | .................... | |
| A | 7405 | hCoV-19/Australia/NSW123/2020\|EPI_ISL_427780\|2020- | .................... | .................... | .................... | .................... | .................... | .................... | |
| A | 7406 | hCoV-19/Australia/NSW159/2020\|EPI_ISL_427781\|2020- | ..................T. | .................... | .................... | .................... | .................... | .................... | 1 |
| A | 7407 | hCoV-19/Australia/NSW124/2020\|EPI_ISL_427782\|2020- | .................... | .................... | .................... | .................... | .................... | .................... | |
| A | 7408 | hCoV-19/Australia/NSW131/2020\|EPI_ISL_427783\|2020- | .................... | .................... | .................... | .................... | .................... | .................... | |
| A | 7409 | hCoV-19/Australia/NSW160/2020\|EPI_ISL_427784\|2020- | .................... | .................... | .................... | .................... | .................... | .................... | |
| A | 7410 | hCoV-19/Australia/NSW125/2020\|EPI_ISL_427785\|2020- | .................... | .................... | .................... | .................... | .................... | .................... | |
| A | 7411 | hCoV-19/Australia/NSW191/2020\|EPI_ISL_427786\|2020- | .................... | .................... | .................... | .................... | .................... | .................... | |
| A | 7412 | hCoV-19/Australia/NSW132/2020\|EPI_ISL_427787\|2020- | .................... | .................... | .................... | .................... | .................... | .................... | |
| A | 7413 | hCoV-19/Australia/NSW161/2020\|EPI_ISL_427788\|2020- | .................... | .................... | .................... | .................... | .................... | .................... | |
| A | 7414 | hCoV-19/Australia/NSW163/2020\|EPI_ISL_427789\|2020- | .................... | .................... | .................... | .................... | .................... | .................... | |
| A | 7415 | hCoV-19/Australia/NSW79/2020\|EPI_ISL_427790\|2020-0 | .................... | .................... | .................... | .................... | .................... | .................... | |
| A | 7416 | hCoV-19/Australia/NSW133/2020\|EPI_ISL_427791\|2020- | .................... | .................... | .................... | .................... | .................... | .................... | |
| A | 7417 | hCoV-19/Australia/NSW204/2020\|EPI_ISL_427792\|2020- | .................... | .................... | .................... | .................... | .................... | .................... | |
| A | 7418 | hCoV-19/Australia/NSW185/2020\|EPI_ISL_427793\|2020- | .................... | .................... | .................... | .................... | .................... | .................... | |
| A | 7419 | hCoV-19/Australia/NSW126/2020\|EPI_ISL_427794\|2020- | .................... | .................... | .................... | .................... | .................... | .................... | |
| A | 7420 | hCoV-19/Australia/NSW139/2020\|EPI_ISL_427795\|2020- | .................... | .................... | .................... | .................... | .................... | .................... | |
| A | 7421 | hCoV-19/Australia/NSW150/2020\|EPI_ISL_427796\|2020- | .................... | .................... | .................... | .................... | .................... | .................... | |
| A | 7422 | hCoV-19/Australia/NSW192/2020\|EPI_ISL_427797\|2020- | .................... | .................... | .................... | .................... | .................... | .................... | |

| | | | | | | | | |
|---|---|---|---|---|---|---|---|---|
| A | 7423 hCoV-19/Australia/NSW193/2020\|EPI_ISL_427798\|2020- | ···················· | ···················· | ····················· | ···················· | ···················· | ···················· | |
| A | 7424 hCoV-19/Australia/NSW140/2020\|EPI_ISL_427799\|2020- | ···················· | ···················· | ····················· | ···················· | ···················· | ···················· | |
| A | 7425 hCoV-19/Australia/QLD09/2020\|EPI_ISL_414414\|2020-0 | ···················· | ···················· | ····················· | ···················· | ···················· | ···················· | |
| A | 7426 hCoV-19/Switzerland/GE-HUG-1402/2020\|EPI_ISL_41570 | ···················· | ···················· | ····················· | ···················· | ···················· | ···················· | |
| A | 7427 hCoV-19/Switzerland/BS-HUG-0914/2020\|EPI_ISL_41570 | ···················· | ···················· | ····················· | ···················· | ···················· | ···················· | |
| A | 7428 hCoV-19/Switzerland/SZ-HUG-1417/2020\|EPI_ISL_41570 | ···················· | ···················· | ····················· | ···················· | ···················· | ···················· | |
| A | 7429 hCoV-19/Switzerland/TI-HUG-2045/2020\|EPI_ISL_41570 | ···················· | ···················· | ····················· | ···················· | ···················· | ···················· | |
| A | 7430 hCoV-19/Switzerland/BE-HUG-2536/2020\|EPI_ISL_41570 | ···················· | ···················· | ····················· | ···················· | ···················· | ···················· | |
| A | 7431 hCoV-19/Switzerland/GE-HUG-4135/2020\|EPI_ISL_41570 | ···················· | ···················· | ····················· | ···················· | ···················· | ···················· | |
| A | 7432 hCoV-19/Switzerland/GE-HUG-06207/2020\|EPI_ISL_4157 | ···················· | ···················· | ····················· | ···················· | ···················· | ···················· | |
| A | 7433 hCoV-19/Switzerland/GE-HUG-6679/2020\|EPI_ISL_41570 | ···················· | ···················· | ····················· | ···················· | ···················· | ···················· | |
| A | 7434 hCoV-19/Switzerland/GE-HUG-4984/2020\|EPI_ISL_41570 | ···················· | ···················A | ····················· | ···················· | ···················· | ···················· | 1 |
| A | 7435 hCoV-19/Russia/StPetersburg-3524/2020\|EPI_ISL_4157 | ···················· | ···················· | ····················· | ···················· | ···················· | ···················· | |
| A | 7436 hCoV-19/Taiwan/CGMH-CGU-03/2020\|EPI_ISL_415741\|202 | ···················· | ···················· | ····················· | ···················· | ···················· | ···················· | |
| A | 7437 hCoV-19/Taiwan/CGMH-CGU-04/2020\|EPI_ISL_415742\|202 | ···················· | ···················· | ····················· | ···················· | ···················· | ···················· | |
| A | 7438 hCoV-19/Taiwan/CGMH-CGU-05/2020\|EPI_ISL_415743\|202 | ···················· | ···················· | ····················· | ···················· | ···················· | ···················· | |
| A | 7439 hCoV-19/Wales/PHWC-23CD6/2020\|EPI_ISL_418103\|2020- | ···················· | ···A················ | ····················· | ···················· | ···················· | ···················· | |
| A | 7440 hCoV-19/Wales/PHWC-23D6A/2020\|EPI_ISL_418110\|2020- | ···················· | ···A················ | ····················· | ···················· | ···················· | ···················· | |
| A | 7441 hCoV-19/Wales/PHWC-23E2B/2020\|EPI_ISL_418117\|2020- | ···················· | ···················· | ····················· | ···················· | ···················· | ···················· | |
| A | 7442 hCoV-19/Wales/PHWC-23E67/2020\|EPI_ISL_418120\|2020- | ···················· | ···A················ | ····················· | ···················· | ···················· | ···················· | |
| A | 7443 hCoV-19/Wales/PHWC-23E85/2020\|EPI_ISL_418122\|2020- | ···················· | ···················· | ····················· | ···················· | ···················· | ···················· | |
| A | 7444 hCoV-19/Wales/PHWC-23FFB/2020\|EPI_ISL_418136\|2020- | ···················· | ···A················ | ····················· | ···················· | ···················· | ···················· | |
| A | 7445 hCoV-19/Wales/PHWC-24006/2020\|EPI_ISL_418137\|2020- | ···················· | ···················· | ····················· | ···················· | ···················· | ···················· | |
| A | 7446 hCoV-19/Wales/PHWC-24024/2020\|EPI_ISL_418139\|2020- | ···················· | ···················· | ····················· | ···················· | ···················· | ···················· | |
| A | 7447 hCoV-19/Wales/PHWC-24033/2020\|EPI_ISL_418140\|2020- | ···················· | ···················· | ····················· | ···················· | ···················· | ···················· | |
| A | 7448 hCoV-19/Wales/PHWC-240D9/2020\|EPI_ISL_418143\|2020- | ···················· | ···A················ | ····················· | ···················· | ···················· | ···················· | |
| A | 7449 hCoV-19/Wales/PHWC-240E8/2020\|EPI_ISL_418144\|2020- | ···················· | ···A················ | ····················· | ···················· | ···················· | ···················· | |
| A | 7450 hCoV-19/Wales/PHWC-240F7/2020\|EPI_ISL_418145\|2020- | ···················· | ···A················ | ····················· | ···················· | ···················· | ···················· | |
| A | 7451 hCoV-19/Wales/PHWC-24121/2020\|EPI_ISL_418147\|2020- | ···················· | ···A················ | ····················· | ···················· | ···················· | ···················· | |
| A | 7452 hCoV-19/Wales/PHWC-2415E/2020\|EPI_ISL_418149\|2020- | ···················· | ···················· | ····················· | ···················· | ···················· | ···················· | |
| A | 7453 hCoV-19/Wales/PHWC-2416D/2020\|EPI_ISL_418150\|2020- | ···················· | ···A················ | ····················· | ···················· | ···················· | ···················· | |
| A | 7454 hCoV-19/Wales/PHWC-2419A/2020\|EPI_ISL_418151\|2020- | ···················· | ···A················ | ····················· | ···················· | ···················· | ···················· | |
| A | 7455 hCoV-19/Wales/PHWC-2422E/2020\|EPI_ISL_418155\|2020- | ···················· | ···A················ | ····················· | ···················· | ···················· | ···················· | |
| A | 7456 hCoV-19/Wales/PHWC-2423D/2020\|EPI_ISL_418156\|2020- | ···················· | ···A················ | ····················· | ···················· | ···················· | ···················· | |
| A | 7457 hCoV-19/Wales/PHWC-24279/2020\|EPI_ISL_418158\|2020- | ···················· | ···A················ | ····················· | ···················· | ···················· | ···················· | |
| A | 7458 hCoV-19/Wales/PHWC-24288/2020\|EPI_ISL_418159\|2020- | ···················· | ···A················ | ····················· | ···················· | ···················· | ···················· | |
| A | 7459 hCoV-19/Wales/PHWC-242C4/2020\|EPI_ISL_418163\|2020- | ···················· | ···A················ | ····················· | ···················· | ···················· | ···················· | |
| A | 7460 hCoV-19/Japan/P1/2020\|EPI_ISL_419296\|2020-03-08 | ···················· | ···················· | ····················· | ···················· | ···················· | ···················· | |
| A | 7461 hCoV-19/Japan/P2-1/2020\|EPI_ISL_419297\|2020-03-10 | ···················· | ···················· | ····················· | ···················· | ···················· | ···················· | |
| A | 7462 hCoV-19/Japan/P2-2/2020\|EPI_ISL_419298\|2020-03-10 | ···················· | ···················· | ····················· | ···················· | ···················· | ···················· | |
| A | 7463 hCoV-19/Japan/P3-1/2020\|EPI_ISL_419299\|2020-03-09 | ···················· | ···················· | ····················· | ···················· | ···················· | ···················· | |
| A | 7464 hCoV-19/Wales/PHWC-243D0/2020\|EPI_ISL_419406\|2020- | ···················· | ···················· | ····················· | ···················· | ···················· | ···················· | |
| A | 7465 hCoV-19/Wales/PHWC-243EF/2020\|EPI_ISL_419407\|2020- | ···················· | ···A················ | ····················· | ···················· | ···················· | ···················· | |
| A | 7466 hCoV-19/Wales/PHWC-24419/2020\|EPI_ISL_419409\|2020- | ···················· | ···A················ | ····················· | ···················· | ···················· | ···················· | |
| A | 7467 hCoV-19/Wales/PHWC-244EC/2020\|EPI_ISL_419420\|2020- | ···················· | ···A················ | ····················· | ···················· | ···················· | ···················· | |
| A | 7468 hCoV-19/Wales/PHWC-244FB/2020\|EPI_ISL_419421\|2020- | ···················· | ···M················ | ····················· | ···················· | ···················· | ···················· | |
| A | 7469 hCoV-19/Wales/PHWC-24507/2020\|EPI_ISL_419422\|2020- | ···················· | ···A················ | ····················· | ···················· | ···················· | ···················· | |
| A | 7470 hCoV-19/Wales/PHWC-24525/2020\|EPI_ISL_419424\|2020- | ···················· | ···A················ | ····················· | ···················· | ···················· | ···················· | |
| A | 7471 hCoV-19/Wales/PHWC-24534/2020\|EPI_ISL_419425\|2020- | ···················· | ···················· | ····················· | ···················· | ···················· | ···················· | |
| A | 7472 hCoV-19/Wales/PHWC-2458F/2020\|EPI_ISL_419428\|2020- | ···················· | ···················· | ····················· | ···················· | ···················· | ···················· | |
| A | 7473 hCoV-19/Wales/PHWC-2465F/2020\|EPI_ISL_419434\|2020- | ···················· | ···················· | ····················· | ···················· | ···················· | ···················· | |
| A | 7474 hCoV-19/Wales/PHWC-2469B/2020\|EPI_ISL_419438\|2020- | ···················· | ···················· | ····················· | ···················· | ···················· | ···················· | |
| A | 7475 hCoV-19/Wales/PHWC-246AA/2020\|EPI_ISL_419439\|2020- | ···················· | ···A················ | ····················· | ···················· | ···················· | ···················· | |
| A | 7476 hCoV-19/Wales/PHWC-246B9/2020\|EPI_ISL_419440\|2020- | ···················· | ···················· | ····················· | ···················· | ···················· | ···················· | |
| A | 7477 hCoV-19/Wales/PHWC-246E6/2020\|EPI_ISL_419443\|2020- | ···················· | ···················· | ····················· | ···················· | ···················· | ···················· | |
| A | 7478 hCoV-19/Wales/PHWC-247F2/2020\|EPI_ISL_419448\|2020- | ···················· | ···A················ | ····················· | ···················· | ···················· | ···················· | |
| A | 7479 hCoV-19/Wales/PHWC-2480E/2020\|EPI_ISL_419449\|2020- | ···················· | ···A················ | ····················· | ···················· | ···················· | ···················· | |
| A | 7480 hCoV-19/Wales/PHWC-248B3/2020\|EPI_ISL_419453\|2020- | ···················· | ···················· | ····················· | ···················· | ···················· | ···················· | |
| A | 7481 hCoV-19/Wales/PHWC-24A35/2020\|EPI_ISL_419472\|2020- | ···················· | ···A················ | ····················· | ···················· | ···················· | ···················· | |
| A | 7482 hCoV-19/Wales/PHWC-24A62/2020\|EPI_ISL_419474\|2020- | ···················· | ···A················ | ····················· | ···················· | ···················· | ···················· | |
| A | 7483 hCoV-19/Wales/PHWC-24BC9/2020\|EPI_ISL_419496\|2020- | ···················· | ···················· | ····················· | ···················· | ···················· | ···················· | |
| A | 7484 hCoV-19/Wales/PHWC-24BE7/2020\|EPI_ISL_419498\|2020- | ···················· | ···A················ | ····················· | ···················· | ···················· | ···················· | |
| A | 7485 hCoV-19/Wales/PHWC-24BF6/2020\|EPI_ISL_419499\|2020- | ···················· | ···A················ | ····················· | ···················· | ···················· | ···················· | |
| A | 7486 hCoV-19/England/SHEF-C0172/2020\|EPI_ISL_420259\|202 | ···················· | ···················· | ····················· | ···················· | ···················· | ···················· | |
| A | 7487 hCoV-19/England/SHEF-C05A3/2020\|EPI_ISL_420279\|202 | ···················· | ···················· | ····················· | ···················· | ···················· | ···················· | |
| A | 7488 hCoV-19/England/SHEF-C06A0/2020\|EPI_ISL_420280\|202 | ···················· | ···················· | ····················· | ···················· | ···················· | ···················· | |
| A | 7489 hCoV-19/England/SHEF-BFFF4/2020\|EPI_ISL_420281\|202 | ···················· | ···················· | ····················· | ···················· | ···················· | ···················· | |
| A | 7490 hCoV-19/England/SHEF-C05C1/2020\|EPI_ISL_420282\|202 | ···················· | ···················· | ····················· | ···················· | ···················· | ···················· | |
| A | 7491 hCoV-19/England/SHEF-C001B/2020\|EPI_ISL_420283\|202 | ···················· | ···················· | ····················· | ···················· | ···················· | ···················· | |
| A | 7492 hCoV-19/England/SHEF-C0576/2020\|EPI_ISL_420284\|202 | ···················· | ···················· | ····················· | ···················· | ···················· | ···················· | |
| A | 7493 hCoV-19/England/SHEF-C060A/2020\|EPI_ISL_420285\|202 | ···················· | ···················· | ····················· | ···················· | ···················· | ···················· | |
| A | 7494 hCoV-19/England/SHEF-C00A2/2020\|EPI_ISL_420286\|202 | ···················· | ···················· | ····················· | ···················· | ···················· | ···················· | |
| A | 7495 hCoV-19/England/SHEF-C0048/2020\|EPI_ISL_420287\|202 | ···················· | ···················· | ····················· | ···················· | ···················· | ···················· | |
| A | 7496 hCoV-19/England/SHEF-C00FD/2020\|EPI_ISL_420288\|202 | ···················· | ···················· | ····················· | ···················· | ···················· | ···················· | |
| A | 7497 hCoV-19/England/SHEF-C0619/2020\|EPI_ISL_420289\|202 | ···················· | ···················· | ····················· | ···················· | ···················· | ···················· | |
| A | 7499 hCoV-19/England/SHEF-C00C0/2020\|EPI_ISL_420291\|202 | ···················· | ···················· | ····················· | ···················· | ···················· | ···················· | |
| A | 7500 hCoV-19/England/SHEF-C0655/2020\|EPI_ISL_420292\|202 | ···················· | ···················· | ····················· | ···················· | ···················· | ···················· | |
| A | 7501 hCoV-19/tiger/USA/NY-040420/2020\|EPI_ISL_420293\|20 | ···················· | ···················· | ····················· | ···················· | ···················· | ···················· | |

| | | | | | | | | |
|---|---|---|---|---|---|---|---|---|
| A | 7502 | hCoV-19/Slovenia/4584/2020\|EPI_ISL_420294\|2020-03- | ···················· | ···················· | ···················· | ···················· | ···················· | ···················· |
| A | 7503 | hCoV-19/Slovenia/12304/2020\|EPI_ISL_420295\|2020-03 | ···················· | ···················· | ···················· | ···················· | ···················· | ···················· |
| A | 7504 | hCoV-19/USA/NY-NYUMC45/2020\|EPI_ISL_420296\|2020-03 | ···················· | ···················· | ···················· | ···················· | ···················· | ···················· |
| A | 7505 | hCoV-19/USA/NY-NYUMC46/2020\|EPI_ISL_420297\|2020-03 | ···················· | ···················· | ···················· | ···················· | ···················· | ···················· |
| A | 7506 | hCoV-19/USA/NY-NYUMC47/2020\|EPI_ISL_420298\|2020-03 | ···················· | ···················· | ···················· | ···················· | ···················· | ···················· |
| A | 7507 | hCoV-19/USA/NY-NYUMC48/2020\|EPI_ISL_420299\|2020-03 | ···················· | ···················· | ···················· | ···················· | ···················· | ···················· |
| A | 7508 | hCoV-19/Belgium/VT-0325143/2020\|EPI_ISL_420400\|202 | ···················· | ···················· | ···················· | ···················· | ···················· | ···················· |
| A | 7509 | hCoV-19/Belgium/PM-0324144/2020\|EPI_ISL_420401\|202 | ···················· | ···················· | ···················· | ···················· | ···················· | ···················· |
| A | 7510 | hCoV-19/Belgium/PC-0325147/2020\|EPI_ISL_420404\|202 | ···················· | ···················· | ···················· | ···················· | ···················· | ···················· |
| A | 7511 | hCoV-19/Belgium/LJ-0325148/2020\|EPI_ISL_420405\|202 | ···················· | ···················· | ···················· | ···················· | ···················· | ···················· |
| A | 7512 | hCoV-19/Belgium/BEG-0324149/2020\|EPI_ISL_420406\|20 | ···················· | ···················· | ···················· | ···················· | ···················· | ···················· |
| A | 7513 | hCoV-19/Belgium/CK-0324150/2020\|EPI_ISL_420407\|202 | ···················· | ···················· | ···················· | ···················· | ···················· | ···················· |
| A | 7514 | hCoV-19/Belgium/HCM-0324151/2020\|EPI_ISL_420408\|20 | ···················· | ···················· | ···················· | ···················· | ···················· | ···················· |
| A | 7515 | hCoV-19/Belgium/ZF-0325152/2020\|EPI_ISL_420409\|202 | ···················· | ···················· | ···················· | ···················· | ···················· | ···················· |
| A | 7516 | hCoV-19/Belgium/VPMV-0325153/2020\|EPI_ISL_420410\|2 | ···················· | ···················· | ···················· | ···················· | ···················· | ···················· |
| A | 7517 | hCoV-19/Belgium/BEG-0323154/2020\|EPI_ISL_420411\|20 | ···················· | ···················· | ···················· | ···················· | ···················· | ···················· |
| A | 7518 | hCoV-19/Belgium/BJ-0324192/2020\|EPI_ISL_420412\|202 | ···················· | ···················· | ···················· | ···················· | ···················· | ···················· |
| A | 7519 | hCoV-19/Belgium/DJMG-0325156/2020\|EPI_ISL_420413\|2 | ···················· | ···················· | ···················· | ···················· | ···················· | ···················· |
| A | 7520 | hCoV-19/Belgium/DRV-0325157/2020\|EPI_ISL_420414\|20 | ···················· | ···················· | ···················· | ···················· | ···················· | ···················· |
| A | 7521 | hCoV-19/Belgium/DCS-0325158/2020\|EPI_ISL_420415\|20 | ···················· | ···················· | ···················· | ···················· | ···················· | ···················· |
| A | 7522 | hCoV-19/Belgium/FMC-0323159/2020\|EPI_ISL_420416\|20 | ···················· | ···················· | ···················· | ···················· | ···················· | ···················· |
| A | 7523 | hCoV-19/Belgium/BCM-0324160/2020\|EPI_ISL_420417\|20 | ···················· | ···················· | ···················· | ···················· | ···················· | ···················· |
| A | 7524 | hCoV-19/Belgium/NIM-0324161/2020\|EPI_ISL_420418\|20 | ···················· | ···················· | ···················· | ···················· | ···················· | ···················· |
| A | 7525 | hCoV-19/Belgium/VLG-0325162/2020\|EPI_ISL_420419\|20 | ···················· | ···················· | ···················· | ···················· | ···················· | ···················· |
| A | 7526 | hCoV-19/Belgium/DCB-0324163/2020\|EPI_ISL_420420\|20 | ···················· | ···················· | ···················· | ···················· | ···················· | ···················· |
| A | 7527 | hCoV-19/Belgium/ML-0324164/2020\|EPI_ISL_420421\|202 | ···················· | ···················· | ···················· | ···················· | ···················· | ···················· |
| A | 7528 | hCoV-19/Belgium/JJC-0325165/2020\|EPI_ISL_420422\|20 | ···················· | ···················· | ···················· | ···················· | ···················· | ···················· |
| A | 7529 | hCoV-19/Belgium/PJM-0325166/2020\|EPI_ISL_420423\|20 | ···················· | ···················· | ···················· | ···················· | ···················· | ···················· |
| A | 7530 | hCoV-19/Belgium/CCM-0324167/2020\|EPI_ISL_420424\|20 | ···················· | ···················· | ···················· | ···················· | ···················· | ···················· |
| A | 7531 | hCoV-19/Belgium/RC-0324168/2020\|EPI_ISL_420425\|202 | ···················· | ···················· | ···················· | ···················· | ···················· | ···················· |
| A | 7532 | hCoV-19/Belgium/NMA-0324169/2020\|EPI_ISL_420426\|20 | ···················· | ···················· | ···················· | ···················· | ···················· | ···················· |
| A | 7533 | hCoV-19/Belgium/STM-0324170/2020\|EPI_ISL_420427\|20 | ···················· | ···················· | ···················· | ···················· | ···················· | ···················· |
| A | 7534 | hCoV-19/Belgium/VM-0325171/2020\|EPI_ISL_420428\|202 | ···················· | ···················· | ···················· | ···················· | ···················· | ···················· |
| A | 7535 | hCoV-19/Belgium/NKMA-0324172/2020\|EPI_ISL_420429\|2 | ···················· | ···················· | ···················· | ···················· | ···················· | ···················· |
| A | 7536 | hCoV-19/Belgium/QL-0324173/2020\|EPI_ISL_420430\|202 | ···················· | ···················· | ···················· | ···················· | ···················· | ···················· |
| A | 7537 | hCoV-19/Belgium/AGR-0324174/2020\|EPI_ISL_420431\|20 | ···················· | ···················· | ···················· | ···················· | ···················· | ···················· |
| A | 7538 | hCoV-19/Belgium/CJM-0323175/2020\|EPI_ISL_420432\|20 | ···················· | ···················· | ···················· | ···················· | ···················· | ···················· |
| A | 7539 | hCoV-19/Belgium/DNS-0324176/2020\|EPI_ISL_420433\|20 | ···················· | ···················· | ···················· | ···················· | ···················· | ···················· |
| A | 7540 | hCoV-19/Belgium/RHA-0324177/2020\|EPI_ISL_420434\|20 | ···················· | ···················· | ···················· | ···················· | ···················· | ···················· |
| A | 7541 | hCoV-19/Belgium/CJE-0325178/2020\|EPI_ISL_420435\|20 | ···················· | ···················· | ···················· | ···················· | ···················· | ···················· |
| A | 7542 | hCoV-19/Belgium/GLA-0324179/2020\|EPI_ISL_420436\|20 | ···················· | ···················· | ···················· | ···················· | ···················· | ···················· |
| A | 7543 | hCoV-19/Belgium/DMM-0325180/2020\|EPI_ISL_420437\|20 | ···················· | ···················· | ···················· | ···················· | ···················· | ···················· |
| A | 7544 | hCoV-19/Belgium/BJ-0325181/2020\|EPI_ISL_420438\|202 | ···················· | ···················· | ···················· | ···················· | ···················· | ···················· |
| A | 7545 | hCoV-19/Belgium/PC-0325182/2020\|EPI_ISL_420439\|202 | ···················· | ···················· | ···················· | ···················· | ···················· | ···················· |
| A | 7546 | hCoV-19/Belgium/BS-0325183/2020\|EPI_ISL_420440\|202 | ···················· | ···················· | ···················· | ···················· | ···················· | ···················· |
| A | 7547 | hCoV-19/Belgium/RA-030664/2020\|EPI_ISL_420441\|2020 | ···················· | ···················· | ···················· | ···················· | ···················· | ···················· |
| A | 7548 | hCoV-19/Belgium/BGM-030444/2020\|EPI_ISL_420442\|202 | ···················· | ···················· | ···················· | ···················· | ···················· | ···················· |
| A | 7549 | hCoV-19/Belgium/RA-030445/2020\|EPI_ISL_420443\|2020 | ···················· | ···················· | ···················· | ···················· | ···················· | ···················· |
| A | 7550 | hCoV-19/Belgium/MF-030546/2020\|EPI_ISL_420444\|2020 | ···················· | ···················· | ···················· | ···················· | ···················· | ···················· |
| A | 7551 | hCoV-19/Belgium/FAE-030948/2020\|EPI_ISL_420445\|202 | ···················· | ···················· | ···················· | ···················· | ···················· | ···················· |
| A | 7552 | hCoV-19/Belgium/DAL-030650/2020\|EPI_ISL_420446\|202 | ···················· | ···················· | ···················· | ···················· | ···················· | ···················· |
| A | 7553 | hCoV-19/Belgium/DJ-030452/2020\|EPI_ISL_420447\|2020 | ···················· | ···················· | ···················· | ···················· | ···················· | ···················· |
| A | 7554 | hCoV-19/Belgium/GSA-030454/2020\|EPI_ISL_420448\|202 | ···················· | ···················· | ···················· | ···················· | ···················· | ···················· |
| A | 7555 | hCoV-19/Belgium/VHC-030655/2020\|EPI_ISL_420449\|202 | ···················· | ···················· | ···················· | ···················· | ···················· | ···················· |
| A | 7556 | hCoV-19/Belgium/KN-030756/2020\|EPI_ISL_420450\|2020 | ···················· | ···················· | ···················· | ···················· | ···················· | ···················· |
| A | 7557 | hCoV-19/Belgium/HC-030760/2020\|EPI_ISL_420451\|2020 | ···················· | ···················· | ···················· | ···················· | ···················· | ···················· |
| A | 7558 | hCoV-19/Belgium/RDC-030961/2020\|EPI_ISL_420452\|202 | ···················· | ···················· | ···················· | ···················· | ···················· | ···················· |
| A | 7559 | hCoV-19/Belgium/VF-030562/2020\|EPI_ISL_420453\|2020 | ···················· | ···················· | ···················· | ···················· | ···················· | ···················· |
| A | 7560 | hCoV-19/Belgium/VJP-030463/2020\|EPI_ISL_420454\|202 | ···················· | ···················· | ···················· | ···················· | ···················· | ···················· |
| A | 7561 | hCoV-19/Australia/WA11/2020\|EPI_ISL_420456\|2020-02 | ···················· | ···················· | ···················· | ···················· | ···················· | ···················· |
| A | 7562 | hCoV-19/England/20114052703/2020\|EPI_ISL_420458\|20 | ···················· | ···················· | ···················· | ···················· | ···················· | ···················· |
| A | 7563 | hCoV-19/England/20120033002/2020\|EPI_ISL_420464\|20 | ···················· | ···················· | ···················· | ···················· | ···················· | ···················· |
| A | 7564 | hCoV-19/England/20126063003/2020\|EPI_ISL_420466\|20 | ··T················· | ···················· | ···················· | ···················· | ···················· | ···················· |
| A | 7565 | hCoV-19/England/20128035202/2020\|EPI_ISL_420477\|20 | ···················· | ···················· | ···················· | ···················· | ···················· | ···················· |
| A | 7566 | hCoV-19/England/20128046202/2020\|EPI_ISL_420478\|20 | ···················· | ···················· | ···················· | ···················· | ···················· | ···················· |
| A | 7567 | hCoV-19/England/20128046302/2020\|EPI_ISL_420479\|20 | ···················· | ···················· | ···················· | ···················· | ···················· | ···················· |
| A | 7568 | hCoV-19/England/20129040102/2020\|EPI_ISL_420488\|20 | ···················· | ···················· | ···················· | ···················· | ···················· | ···················· |
| A | 7570 | hCoV-19/USA/NY-NYUMC87/2020\|EPI_ISL_421588\|2020-03 | ···················· | ···················· | ···················· | ···················· | ···················· | ···················· |
| A | 7571 | hCoV-19/USA/NY-NYUMC88/2020\|EPI_ISL_421589\|2020-03 | ···················· | ···················· | ···················· | ···················· | ···················· | ···················· |
| A | 7572 | hCoV-19/USA/NY-NYUMC94/2020\|EPI_ISL_421706\|2020-03 | ···················· | ···················· | ···················· | ···················· | ···················· | ···················· |
| A | 7573 | hCoV-19/USA/NY-NYUMC95/2020\|EPI_ISL_421707\|2020-03 | ···················· | ···················· | ···················· | ···················· | ···················· | ···················· |
| A | 7574 | hCoV-19/USA/NY-NYUMC97/2020\|EPI_ISL_421709\|2020-03 | ···················· | ···················· | ···················· | ···················· | ···················· | ···················· |
| A | 7575 | hCoV-19/USA/NY-NYUMC98/2020\|EPI_ISL_421710\|2020-03 | ···················· | ···················· | ···················· | ···················· | ···················· | ···················· |
| A | 7576 | hCoV-19/USA/NY-NYUMC99/2020\|EPI_ISL_421711\|2020-03 | ···················· | ···················· | ···················· | ···················· | ···················· | ···················· |
| A | 7577 | hCoV-19/USA/NY-NYUMC100/2020\|EPI_ISL_421712\|2020-0 | ···················· | ···················· | ···················· | ···················· | ···················· | ···················· |
| A | 7578 | hCoV-19/USA/NY-NYUMC102/2020\|EPI_ISL_421714\|2020-0 | ···················· | ···················· | ···················· | ···················· | ···················· | ···················· |
| A | 7579 | hCoV-19/USA/NY-NYUMC103/2020\|EPI_ISL_421715\|2020-0 | ···················· | ···················· | ···················· | ···················· | ···················· | ···················· |
| A | 7580 | hCoV-19/USA/NY-NYUMC105/2020\|EPI_ISL_421717\|2020-0 | ···················· | ···················· | ···················· | ···················· | ···················· | ···················· |

A 7581 hCoV-19/USA/NJ-NYUMC106/2020|EPI_ISL_421718|2020-0 .................... .................... .................... .................... .................... ....................
A 7582 hCoV-19/USA/NY-NYUMC107/2020|EPI_ISL_421719|2020-0 .................... .................... .................... .................... .................... ....................
A 7583 hCoV-19/USA/NY-NYUMC121/2020|EPI_ISL_421733|2020-0 .................... .................... .................... .................... .................... ....................
A 7584 hCoV-19/Luxembourg/LNS3307371/2020|EPI_ISL_421735| .................... .................... .................... .................... .................... ....................
A 7585 hCoV-19/Luxembourg/LNS2334563/2020|EPI_ISL_421737| .................... .................... .................... .................... .................... ....................
A 7586 hCoV-19/Luxembourg/LNS3404815/2020|EPI_ISL_421738| .................... .................... .................... .................... .................... ....................
A 7587 hCoV-19/Luxembourg/LNS1644489/2020|EPI_ISL_421740| .................... .................... .................... .................... .................... ....................
A 7588 hCoV-19/Luxembourg/LNS6689955/2020|EPI_ISL_421741| .................... .................... .................... .................... .................... ....................
A 7589 hCoV-19/Luxembourg/LNS6500589/2020|EPI_ISL_421742| .................... .................... .................... .................... .................... ....................
A 7590 hCoV-19/Luxembourg/LNS0272441/2020|EPI_ISL_421743| .................... .................... .................... .................... .................... ....................
A 7591 hCoV-19/Luxembourg/LNS7074589/2020|EPI_ISL_421744| .................... .................... .................... .................... .................... ....................
A 7592 hCoV-19/Luxembourg/LNS2299410/2020|EPI_ISL_421745| .................... .................... .................... .................... .................... ....................
A 7593 hCoV-19/Luxembourg/LNS0819394/2020|EPI_ISL_421746| .................... .................... .................... .................... .................... ....................
A 7594 hCoV-19/Luxembourg/LNS3770609/2020|EPI_ISL_421747| .................... .................... .................... .................... .................... ....................
A 7595 hCoV-19/Luxembourg/LNS3052946/2020|EPI_ISL_421748| .................... .................... .................... .................... .................... ....................
A 7596 hCoV-19/Luxembourg/LNS3035957/2020|EPI_ISL_421750| .................... .................... .................... .................... .................... ....................
A 7597 hCoV-19/Luxembourg/LNS4569788/2020|EPI_ISL_421752| .................... .................... .................... .................... .................... ....................
A 7598 hCoV-19/Luxembourg/LNS7991123/2020|EPI_ISL_421753| .................... .................... .................... .................... .................... ....................
A 7599 hCoV-19/Luxembourg/LNS0814536/2020|EPI_ISL_421754| .................... .................... .................... .................... .................... ....................
A 7600 hCoV-19/Luxembourg/LNS6897699/2020|EPI_ISL_421755| .................... .................... .................... .................... .................... ....................
A 7601 hCoV-19/Luxembourg/LNS8124566/2020|EPI_ISL_421756| .................... .................... .................... .................... .................... ....................
A 7602 hCoV-19/Luxembourg/LNS7914820/2020|EPI_ISL_421757| .................... .................... .................... .................... .................... ....................
A 7603 hCoV-19/Luxembourg/LNS4235576/2020|EPI_ISL_421759| .................... .................... .................... .................... .................... ....................
A 7604 hCoV-19/Luxembourg/LNS2458673/2020|EPI_ISL_421760| .................... .................... .................... .................... .................... ....................
A 7605 hCoV-19/Luxembourg/LNS8602041/2020|EPI_ISL_421761| .................... .................... .................... .................... .................... ....................
A 7606 hCoV-19/Luxembourg/LNS9510711/2020|EPI_ISL_421762| .................... .................... .................... .................... .................... ....................
A 7607 hCoV-19/England/201360017/2020|EPI_ISL_421768|2020 .................... .................... .................... .................... .................... ....................
A 7609 hCoV-19/England/201360185/2020|EPI_ISL_421771|2020 .................... .................... .................... .................... .................... ....................
A 7610 hCoV-19/England/201360187/2020|EPI_ISL_421772|2020 .................... .................... .................... .................... .................... ....................
A 7611 hCoV-19/England/201360289/2020|EPI_ISL_421773|2020 .................... .................... .................... .................... .................... ....................
A 7612 hCoV-19/England/201360295/2020|EPI_ISL_421774|2020 .................... .................... .................... .................... .................... ....................
A 7613 hCoV-19/England/201360297/2020|EPI_ISL_421776|2020 .................... .................... .................... .................... .................... ....................
A 7614 hCoV-19/England/201360298/2020|EPI_ISL_421777|2020 .................... .................... .................... .................... .................... ....................
A 7615 hCoV-19/England/201360299/2020|EPI_ISL_421778|2020 .................... .................... .................... .................... .................... ....................
A 7616 hCoV-19/England/201360300/2020|EPI_ISL_421779|2020 .................... .................... .................... .................... .................... ....................
A 7617 hCoV-19/England/201360492/2020|EPI_ISL_421780|2020 .................... .................... .................... .................... .................... ....................
A 7618 hCoV-19/England/201360499/2020|EPI_ISL_421781|2020 .................... .................... .................... .................... .................... ....................
A 7619 hCoV-19/England/201360506/2020|EPI_ISL_421782|2020 .................... .................... .................... .................... .................... ....................
A 7620 hCoV-19/England/201360508/2020|EPI_ISL_421783|2020 .................... .................... .................... .................... .................... ....................
A 7621 hCoV-19/England/201361007/2020|EPI_ISL_421784|2020 .................... .................... .................... .................... .................... ....................
A 7622 hCoV-19/England/201361008/2020|EPI_ISL_421785|2020 .................... .................... .................... .................... .................... ....................
A 7623 hCoV-19/England/201361012/2020|EPI_ISL_421787|2020 .................... .................... .................... .................... .................... ....................
A 7625 hCoV-19/England/201361022/2020|EPI_ISL_421790|2020 .................... .................... .................... .................... .................... ....................
A 7626 hCoV-19/England/201361040/2020|EPI_ISL_421791|2020 .................... .................... .................... .................... .................... ....................
A 7628 hCoV-19/England/201380024/2020|EPI_ISL_421794|2020 .................... .................... .................... .................... .................... ....................
A 7629 hCoV-19/England/201380025/2020|EPI_ISL_421795|2020 .................... .................... .................... .................... .................... ....................
A 7630 hCoV-19/England/201380028/2020|EPI_ISL_421796|2020 .................... .................... .................... .................... .................... ....................
A 7631 hCoV-19/England/201380030/2020|EPI_ISL_421797|2020 .................... .................... .................... .................... .................... ....................
A 7632 hCoV-19/England/201380034/2020|EPI_ISL_421798|2020 .................... .................... .................... .................... .................... ....................
A 7634 hCoV-19/England/CAMB-73A46/2020|EPI_ISL_425270|202 .................... .................... .................... .................... .................... ....................
A 7636 hCoV-19/England/CAMB-73ABF/2020|EPI_ISL_425275|202 .................... .................... .................... .................... .................... ....................
A 7637 hCoV-19/England/CAMB-73ACE/2020|EPI_ISL_425276|202 .................... .................... .................... .................... .................... ....................
A 7638 hCoV-19/England/CAMB-73ADD/2020|EPI_ISL_425277|202 .................... .................... .................... .................... .................... ....................
A 7639 hCoV-19/England/CAMB-73AEC/2020|EPI_ISL_425278|202 .................... .................... .................... .................... .................... ....................
A 7640 hCoV-19/England/CAMB-74359/2020|EPI_ISL_425280|202 .................... .................... .................... .................... .................... ....................
A 7641 hCoV-19/England/CAMB-74368/2020|EPI_ISL_425281|202 .................... .................... .................... .................... .................... ....................
A 7642 hCoV-19/England/CAMB-74386/2020|EPI_ISL_425283|202 .................... .................... .................... .................... .................... ....................
A 7643 hCoV-19/England/CAMB-743E0/2020|EPI_ISL_425286|202 .................... .................... .................... .................... .................... ....................
A 7644 hCoV-19/England/CAMB-743FF/2020|EPI_ISL_425287|202 .................... .................... .................... .................... .................... ....................
A 7645 hCoV-19/England/CAMB-7440B/2020|EPI_ISL_425288|202 .................... .................... .................... .................... .................... ....................
A 7646 hCoV-19/England/CAMB-74438/2020|EPI_ISL_425290|202 .................... .................... .................... .................... .................... ....................
A 7647 hCoV-19/England/CAMB-74465/2020|EPI_ISL_425292|202 .................... .................... .................... .................... .................... ....................
A 7648 hCoV-19/England/CAMB-74474/2020|EPI_ISL_425293|202 .................... .................... .................... .................... .................... ....................
A 7649 hCoV-19/England/CAMB-74483/2020|EPI_ISL_425294|202 .................... .................... .................... .................... .................... ....................
A 7650 hCoV-19/England/CAMB-74492/2020|EPI_ISL_425295|202 .................... .................... .................... .................... .................... ....................
A 7651 hCoV-19/England/CAMB-744A1/2020|EPI_ISL_425296|202 .................... .................... .................... .................... .................... ....................
A 7652 hCoV-19/England/CAMB-744CF/2020|EPI_ISL_425298|202 .................... .................... .................... .................... .................... ....................
A 7653 hCoV-19/Australia/NSW137/2020|EPI_ISL_427700|2020- .................... .................... .................... .................... .................... ....................
A 7654 hCoV-19/Australia/NSW180/2020|EPI_ISL_427701|2020- .................... .................... .................... .................... .................... ....................
A 7655 hCoV-19/Australia/NSW88/2020|EPI_ISL_427702|2020-0 .................... .................... .................... .................... .................... ....................
A 7656 hCoV-19/Australia/NSW75/2020|EPI_ISL_427703|2020-0 .................... .................... .................... .................... .................... ....................
A 7657 hCoV-19/Australia/NSW76/2020|EPI_ISL_427704|2020-0 .................... .................... .................... .................... .................... ....................
A 7658 hCoV-19/Australia/NSW69/2020|EPI_ISL_427705|2020-0 .................... .................... .................... .................... .................... ....................
A 7659 hCoV-19/Australia/NSW74/2020|EPI_ISL_427706|2020-0 .................... .................... .................... .................... .................... ....................
A 7660 hCoV-19/Australia/NSW198/2020|EPI_ISL_427707|2020- .................... .................... .................... .................... .................... ....................
A 7661 hCoV-19/Australia/NSW145/2020|EPI_ISL_427708|2020- .................... .................... .................... .................... .................... ....................
A 7662 hCoV-19/Australia/NSW181/2020|EPI_ISL_427709|2020- .................... .................... .................... .................... .................... ....................
A 7663 hCoV-19/Australia/NSW201/2020|EPI_ISL_427710|2020- .................... .................... .................... .................... .................... ....................

A 7664 hCoV-19/Australia/NSW83/2020|EPI_ISL_427711|2020-0 ···················· ···················· ····················· ···················· ···················· ····················
A 7665 hCoV-19/Australia/NSW91/2020|EPI_ISL_427712|2020-0 ···················· ···················· ····················· ···················· ···················· ····················
A 7666 hCoV-19/Australia/NSW182/2020|EPI_ISL_427713|2020- ···················· ···················· ····················· ···················· ···················· ····················
A 7667 hCoV-19/Australia/NSW183/2020|EPI_ISL_427714|2020- ···················· ···················· ····················· ···················· ···················· ····················
A 7668 hCoV-19/Australia/NSW164/2020|EPI_ISL_427715|2020- ···················· ···················· ····················· ···················· ···················· ····················
A 7669 hCoV-19/Australia/NSW94/2020|EPI_ISL_427716|2020-0 ···················· ···················· ····················· ···················· ···················· ····················
A 7670 hCoV-19/Australia/NSW101/2020|EPI_ISL_427717|2020- ···················· ···················· ····················· ···················· ···················· ····················
A 7671 hCoV-19/Australia/NSW85/2020|EPI_ISL_427718|2020-0 ···················· ···················· ····················· ···················· ···················· ····················
A 7672 hCoV-19/Australia/NSW156/2020|EPI_ISL_427719|2020- ···················· ···················· ····················· ···················· ···················· ····················
A 7673 hCoV-19/Australia/NSW81/2020|EPI_ISL_427720|2020-0 ···················· ···················· ····················· ···················· ···················· ····················
A 7674 hCoV-19/Australia/NSW151/2020|EPI_ISL_427721|2020- ···················· ···················· ····················· ···················· ···················· ····················
A 7675 hCoV-19/Australia/NSW100/2020|EPI_ISL_427722|2020- ···················· ···················· ····················· ···················· ···················· ····················
A 7676 hCoV-19/Australia/NSW84/2020|EPI_ISL_427723|2020-0 ···················· ···················· ····················· ···················· ···················· ····················
A 7677 hCoV-19/Australia/NSW114/2020|EPI_ISL_427724|2020- ···················· ···················· ····················· ···················· ···················· ····················
A 7678 hCoV-19/Australia/NSW86/2020|EPI_ISL_427725|2020-0 ···················· ···················· ····················· ···················· ···················· ····················
A 7679 hCoV-19/Germany/BY-ChVir-1530/2020|EPI_ISL_450210| ···················· ···················· ····················· ···················· ···················· ····················
A 7680 hCoV-19/Germany/BY-ChVir-1536/2020|EPI_ISL_450211| ···················· ···················· ····················· ···················· ···················· ····················
A 7681 hCoV-19/Australia/NSW06/2020|EPI_ISL_413213|2020-0 ···················· ···················· ····················· ···················· ···················· ····················
A 7682 hCoV-19/Australia/NSW07/2020|EPI_ISL_413214|2020-0 ···················· ···················· ····················· ···················· ···················· ····················
A 7683 hCoV-19/Scotland/CVR01/2020|EPI_ISL_413221|2020-02 ···················· ···················· ····················· ···················· ···················· ····················
A 7684 hCoV-19/England/Sheff01/2020|EPI_ISL_414500|2020-0 ···················· ···················· ····················· ···················· ···················· ····················
A 7685 hCoV-19/England/Sheff02/2020|EPI_ISL_414501|2020-0 ···················· ···················· ····················· ···················· ···················· ····················
A 7686 hCoV-19/Germany/NW-HHU-06/2020|EPI_ISL_414505|2020 ···················· ···················· ····················· ···················· ···················· ····················
A 7687 hCoV-19/Germany/NW-HHU-08/2020|EPI_ISL_414508|2020 ···················· ···················· ····················· ···················· ···················· ····················
A 7688 hCoV-19/Germany/NW-HHU-09/2020|EPI_ISL_414509|2020 ···················· ···················· ····················· ···················· ···················· ····················
A 7689 hCoV-19/Shanghai/SH01/2020|EPI_ISL_414510|2020-02- ···················· ···················· ····················· ···················· ···················· ····················
A 7690 hCoV-19/Japan/TKYE6182/2020|EPI_ISL_414511|2020-01 ···················· ···················· ····················· ···················· ···················· ····················
A 7691 hCoV-19/Netherlands/NoordBrabant_28/2020|EPI_ISL_4 ···················· ···················· ····················· ···················· ···················· ····················
A 7692 hCoV-19/Netherlands/NoordBrabant_30/2020|EPI_ISL_4 ···················· ···················· ····················· ···················· ···················· ····················
A 7693 hCoV-19/Netherlands/NoordBrabant_32/2020|EPI_ISL_4 ···················· ···················· ····················· ···················· ···················· ····················
A 7694 hCoV-19/Netherlands/NoordBrabant_33/2020|EPI_ISL_4 ···················· ···················· ····················· ···················· ···················· ····················
A 7695 hCoV-19/Netherlands/NoordBrabant_34/2020|EPI_ISL_4 ···················· ···················· ···············C····· ···················· ···················· ····················
A 7696 hCoV-19/Netherlands/NoordBrabant_35/2020|EPI_ISL_4 ···················· ···················· ····················· ···················· ···················· ····················
A 7697 hCoV-19/Netherlands/NoordBrabant_36/2020|EPI_ISL_4 ···················· ···················· ····················· ···················· ···················· ····················
A 7698 hCoV-19/Netherlands/NoordBrabant_39/2020|EPI_ISL_4 ···················· ···················· ····················· ···················· ···················· ····················
A 7699 hCoV-19/Netherlands/NoordHolland_2/2020|EPI_ISL_41 ···················· ···················· ····················· ···················· ···················· ····················
A 7700 hCoV-19/Netherlands/Utrecht_47/2020|EPI_ISL_414551 ···················· ···················· ····················· ···················· ···················· ····················
A 7701 hCoV-19/Netherlands/Utrecht_46/2020|EPI_ISL_414552 ···················· ···················· ····················· ···················· ···················· ····················
A 7702 hCoV-19/Netherlands/Utrecht_15/2020|EPI_ISL_414554 ···················· ···················· ····················· ···················· ···················· ····················
A 7703 hCoV-19/Netherlands/Utrecht_16/2020|EPI_ISL_414555 ···················· ···················· ····················· ···················· ···················· ····················
A 7704 hCoV-19/Netherlands/ZuidHolland_2/2020|EPI_ISL_414 ···················· ···················· ····················· ···················· ···················· ····················
A 7705 hCoV-19/Netherlands/ZuidHolland_15/2020|EPI_ISL_41 ···················· ···················· ····················· ···················· ···················· ····················
A 7706 hCoV-19/Netherlands/ZuidHolland_16/2020|EPI_ISL_41 ···················· ···················· ····················· ···················· ···················· ····················
A 7707 hCoV-19/Netherlands/ZuidHolland_17/2020|EPI_ISL_41 ···················· ···················· ····················· ···················· ···················· ····················
A 7708 hCoV-19/Netherlands/ZuidHolland_18/2020|EPI_ISL_41 ···················· ···················· ····················· ···················· ···················· ····················
A 7709 hCoV-19/Netherlands/ZuidHolland_19/2020|EPI_ISL_41 ···················· ···················· ····················· ···················· ···················· ····················
A 7710 hCoV-19/Netherlands/ZuidHolland_20/2020|EPI_ISL_41 ···················· ···················· ····················· ···················· ···················· ····················
A 7711 hCoV-19/Netherlands/ZuidHolland_21/2020|EPI_ISL_41 ···················· ···················· ····················· ···················· ···················· ····················
A 7712 hCoV-19/Netherlands/ZuidHolland_22/2020|EPI_ISL_41 ···················· ···················· ····················· ···················· ···················· ····················
A 7713 hCoV-19/Netherlands/ZuidHolland_24/2020|EPI_ISL_41 ···················· ···················· ····················· ···················· ···················· ····················
A 7714 hCoV-19/Hong Kong/VM20002582-2/2020|EPI_ISL_414569 ···················· ···················· ····················· ···················· ···················· ····················
A 7715 hCoV-19/Hong Kong/VM20002849/2020|EPI_ISL_414571|2 ···················· ···················· ····················· ···················· ···················· ····················
A 7716 hCoV-19/Chile/ML-ISPCH-1/2020|EPI_ISL_414577|2020- ···················· ···················· ····················· ···················· ···················· ····················
A 7717 hCoV-19/Chile/ML-ISPCH-2/2020|EPI_ISL_414578|2020- ···················· ···················· ····················· ···················· ···················· ····················
A 7718 hCoV-19/Chile/RM-ISPCH-1/2020|EPI_ISL_414579|2020- ···················· ···················· ····················· ···················· ···················· ····················
A 7719 hCoV-19/Chile/RM-ISPCH-2/2020|EPI_ISL_414580|2020- ···················· ···················· ····················· ···················· ···················· ····················
A 7720 hCoV-19/Ireland/LK-NVRL-19934/2020|EPI_ISL_414586| ···················· ···················· ····················· ···················· ···················· ····················
A 7721 hCoV-19/Ireland/LK-NVRL-19935/2020|EPI_ISL_414587| ···················· ···················· ····················· ···················· ···················· ····················
A 7722 hCoV-19/USA/MN2-MDH2/2020|EPI_ISL_414589|2020-03-0 ···················· ···················· ····················· ···················· ···················· ····················
A 7723 hCoV-19/USA/MN3-MDH3/2020|EPI_ISL_414590|2020-03-0 ···················· ···················· ····················· ···················· ···················· ···············A····
A 7724 hCoV-19/USA/WA-UW22/2020|EPI_ISL_414591|2020-03-06 ···················· ···················· ····················· ···················· ···················· ····················
A 7725 hCoV-19/USA/WA-UW23/2020|EPI_ISL_414592|2020-03-06 ···················· ···················· ····················· ···················· ···················· ····················
A 7726 hCoV-19/USA/WA-UW24/2020|EPI_ISL_414593|2020-03-05 ···················· ···················· ····················· ···················· ···················· ····················
A 7727 hCoV-19/USA/WA-UW25/2020|EPI_ISL_414594|2020-03-05 ···················· ···················· ····················· ···················· ···················· ····················
A 7728 hCoV-19/USA/WA-UW26/2020|EPI_ISL_414595|2020-03-05 ···················· ···················· ····················· ···················· ···················· ····················
A 7729 hCoV-19/USA/WA-UW27/2020|EPI_ISL_414596|2020-03-04 ···················· ···················· ····················· ···················· ···················· ····················
A 7730 hCoV-19/USA/WA-UW28/2020|EPI_ISL_414597|2020-03-04 ···················· ···················· ····················· ···················· ···················· ····················
A 7731 hCoV-19/Spain/VC-FISABIO-3/2020|EPI_ISL_414598|202 ···················· ···················· ····················· ···················· ···················· ····················
A 7732 hCoV-19/USA/WA-UW275/2020|EPI_ISL_418060|2020-03-1 ···················· ···················· ····················· ···················· ···················· ····················
A 7733 hCoV-19/USA/UN-UW276/2020|EPI_ISL_418061|2020-03-1 ···················· ···················· ····················· ···················· ···················· ····················
A 7734 hCoV-19/USA/WA-UW277/2020|EPI_ISL_418062|2020-03-1 ···················· ···················· ····················· ···················· ···················· ····················
A 7735 hCoV-19/USA/WA-UW279/2020|EPI_ISL_418064|2020-03-1 ···················· ···················· ····················· ···················· ···················· ····················
A 7736 hCoV-19/USA/WA-UW282/2020|EPI_ISL_418067|2020-03-1 ···················· ···················· ····················· ···················· ···················· ····················
A 7737 hCoV-19/USA/WA-UW284/2020|EPI_ISL_418069|2020-03-1 ···················· ···················· ····················· ···················· ···················· ····················
A 7738 hCoV-19/USA/WA-UW285/2020|EPI_ISL_418070|2020-03-1 ···················· ···················· ····················· ···················· ···················· ····················
A 7739 hCoV-19/USA/WA-UW286/2020|EPI_ISL_418071|2020-03-1 ···················· ···················· ····················· ···················· ···················· ····················
A 7740 hCoV-19/USA/WA-UW287/2020|EPI_ISL_418072|2020-03-1 ···················· ···················· ····················· ···················· ···················· ····················
A 7741 hCoV-19/USA/WA-UW288/2020|EPI_ISL_418073|2020-03-1 ···················· ···················· ····················· ···················· ···················· ····················

| | | | | | | | | |
|---|---|---|---|---|---|---|---|---|
| A | 7742 hCoV-19/USA/WA-UW289/2020\|EPI_ISL_418074\|2020-03-1 | ···················· | ···················· | ···················· | ···················· | ···················· | ···················· | |
| A | 7743 hCoV-19/USA/WA-UW290/2020\|EPI_ISL_418075\|2020-03-1 | ···················· | ···················· | ···················· | ···················· | ···················· | ···················· | |
| A | 7744 hCoV-19/USA/WA-UW291/2020\|EPI_ISL_418076\|2020-03-1 | ···················· | ···················· | ···················· | ···················· | ···················· | ···················· | |
| A | 7745 hCoV-19/USA/WA-UW292/2020\|EPI_ISL_418077\|2020-03-1 | ···················· | ···················· | ···················· | ···················· | ···················· | ···················· | |
| A | 7746 hCoV-19/USA/WA-UW293/2020\|EPI_ISL_418078\|2020-03-1 | ···················· | ···················· | ···················· | ···················· | ···················· | ···················· | |
| A | 7747 hCoV-19/USA/WA-UW294/2020\|EPI_ISL_418079\|2020-03-1 | ···················· | ···················· | ···················· | ···················· | ···················· | ···················· | |
| A | 7748 hCoV-19/USA/WA-UW295/2020\|EPI_ISL_418080\|2020-03-1 | ···················· | ···················· | ···················· | ···················· | ···················· | ···················· | |
| A | 7749 hCoV-19/USA/WA-UW296/2020\|EPI_ISL_418081\|2020-03-1 | ···················· | ···················· | ···················· | ···················· | ···················· | ···················· | |
| A | 7750 hCoV-19/USA/WA-UW297/2020\|EPI_ISL_418082\|2020-03-1 | ···················· | ···················· | ···················· | ···················· | ···················· | ···················· | |
| A | 7751 hCoV-19/Wales/PHWC-23B60/2020\|EPI_ISL_418093\|2020- | ···················· | ···················· | ···················· | ···················· | ···················· | ···················· | |
| A | 7752 hCoV-19/Wales/PHWC-23BAC/2020\|EPI_ISL_418096\|2020- | ···················· | ···················· | ···················· | ···················· | ···················· | ···················· | |
| A | 7753 hCoV-19/USA/MN62-MDH62/2020\|EPI_ISL_419390\|2020-03 | ···················· | ···················· | ···················· | ···················· | ···················· | ···················· | |
| A | 7754 hCoV-19/USA/MN63-MDH63/2020\|EPI_ISL_419391\|2020-03 | ···················· | ···················· | ···················· | ···················· | ···················· | ···················· | |
| A | 7755 hCoV-19/USA/MN64-MDH64/2020\|EPI_ISL_419392\|2020-03 | ···················· | ···················· | ···················· | ···················· | ···················· | ···················· | |
| A | 7756 hCoV-19/USA/MN65-MDH65/2020\|EPI_ISL_419393\|2020-03 | ···················· | ···················· | ···················· | ···················· | ···················· | ···················· | |
| A | 7757 hCoV-19/USA/MN66-MDH66/2020\|EPI_ISL_419394\|2020-03 | ···················· | ···················· | ···················· | ···················· | ···················· | ···················· | |
| A | 7758 hCoV-19/USA/MN67-MDH67/2020\|EPI_ISL_419395\|2020-03 | ···················· | ···················· | ···················· | ···················· | ···················· | ···················· | |
| A | 7759 hCoV-19/USA/MN68-MDH68/2020\|EPI_ISL_419396\|2020-03 | ···················· | ···················· | ···················· | ···················· | ···················· | ···················· | |
| A | 7760 hCoV-19/USA/MN69-MDH69/2020\|EPI_ISL_419397\|2020-03 | ···················· | ···················· | ···················· | ···················· | ···················· | ···················· | |
| A | 7761 hCoV-19/Wales/PHWC-24349/2020\|EPI_ISL_419399\|2020- | ···················· | ···················· | ···················· | ···················· | ···················· | ···················· | |
| A | 7762 hCoV-19/Wales/PHWC-24455/2020\|EPI_ISL_419412\|2020- | ···················· | ····A··············· | ···················· | ···················· | ···················· | ···················· | |
| A | 7763 hCoV-19/Wales/PHWC-244CE/2020\|EPI_ISL_419418\|2020- | ···················· | ····A··············· | ···················· | ···················· | ···················· | ···················· | |
| A | 7764 hCoV-19/Belgium/MSE-0324121/2020\|EPI_ISL_420378\|20 | ···················· | ···················· | ···················· | ···················· | ···················· | ···················· | |
| A | 7765 hCoV-19/Belgium/RGE-0325122/2020\|EPI_ISL_420379\|20 | ···················· | ···················· | ···················· | ···················· | ···················· | ···················· | |
| A | 7766 hCoV-19/Belgium/DPG-0325123/2020\|EPI_ISL_420380\|20 | ···················· | ···················· | ···················· | ···················· | ···················· | ···················· | |
| A | 7767 hCoV-19/Belgium/FFJ-0324124/2020\|EPI_ISL_420381\|20 | ···················· | ···················· | ···················· | ···················· | ···················· | ···················· | |
| A | 7768 hCoV-19/Belgium/UNJP-0324125/2020\|EPI_ISL_420382\|2 | ···················· | ···················· | ···················· | ···················· | ···················· | ···················· | |
| A | 7769 hCoV-19/Belgium/NJC-0324126/2020\|EPI_ISL_420383\|20 | ···················· | ···················· | ···················· | ···················· | ···················· | ···················· | |
| A | 7770 hCoV-19/Belgium/NML-0325127/2020\|EPI_ISL_420384\|20 | ···················· | ···················· | ···················· | ···················· | ···················· | ···················· | |
| A | 7771 hCoV-19/Belgium/EC-0325128/2020\|EPI_ISL_420385\|202 | ···················· | ···················· | ···················· | ···················· | ···················· | ···················· | |
| A | 7772 hCoV-19/Belgium/MC-0323129/2020\|EPI_ISL_420386\|202 | ···················· | ···················· | ···················· | ···················· | ···················· | ···················· | |
| A | 7773 hCoV-19/Belgium/MNL-0324130/2020\|EPI_ISL_420387\|20 | ···················· | ···················· | ···················· | ···················· | ···················· | ···················· | |
| A | 7774 hCoV-19/Belgium/MSE-0324131/2020\|EPI_ISL_420388\|20 | ···················· | ···················· | ···················· | ···················· | ···················· | ···················· | |
| A | 7775 hCoV-19/Belgium/GMJ-0324132/2020\|EPI_ISL_420389\|20 | ···················· | ···················· | ···················· | ···················· | ···················· | ···················· | |
| A | 7776 hCoV-19/Belgium/LV-0324133/2020\|EPI_ISL_420390\|202 | ···················· | ···················· | ···················· | ···················· | ···················· | ···················· | |
| A | 7777 hCoV-19/Belgium/MS-0324134/2020\|EPI_ISL_420391\|202 | ···················· | ···················· | ···················· | ···················· | ···················· | ···················· | |
| A | 7778 hCoV-19/Belgium/DME-0324135/2020\|EPI_ISL_420392\|20 | ···················· | ···················· | ···················· | ···················· | ···················· | ···················· | |
| A | 7779 hCoV-19/Belgium/VWJM-0324136/2020\|EPI_ISL_420393\|2 | ···················· | ···················· | ···················· | ···················· | ···················· | ···················· | |
| A | 7780 hCoV-19/Belgium/OS-0325137/2020\|EPI_ISL_420394\|202 | ···················· | ···················· | ···················· | ···················· | ···················· | ···················· | |
| A | 7781 hCoV-19/Belgium/RS-0324138/2020\|EPI_ISL_420395\|202 | ···················· | ···················· | ···················· | ···················· | ···················· | ···················· | |
| A | 7782 hCoV-19/Belgium/GM-0323139/2020\|EPI_ISL_420396\|202 | ···················· | ···················· | ···················· | ···················· | ···················· | ···················· | |
| A | 7783 hCoV-19/Belgium/DB-0323140/2020\|EPI_ISL_420397\|202 | ···················· | ···················· | ···················· | ···················· | ···················· | ···················· | |
| A | 7784 hCoV-19/Belgium/NA-0323141/2020\|EPI_ISL_420398\|202 | ···················· | ···················· | ···················· | ···················· | ···················· | ···················· | |
| A | 7785 hCoV-19/Belgium/MAC-0324142/2020\|EPI_ISL_420399\|20 | ···················· | ···················· | ···················· | ···················· | ···················· | ···················· | |
| A | 7786 hCoV-19/Belgium/GJJ-0325145/2020\|EPI_ISL_420402\|20 | ···················· | ···················· | ···················· | ···················· | ···················· | ···················· | |
| A | 7787 hCoV-19/Belgium/DVG-0325146/2020\|EPI_ISL_420403\|20 | ···················· | ···················· | ···················· | ···················· | ···················· | ···················· | |
| A | 7788 hCoV-19/USA/NY-NYUMC108/2020\|EPI_ISL_421720\|2020-0 | ···················· | ···················· | ···················· | ···················· | ···················· | ···················· | |
| A | 7789 hCoV-19/USA/NY-NYUMC109/2020\|EPI_ISL_421721\|2020-0 | ···················· | ···················· | ···················· | ···················· | ···················· | ···················· | |
| A | 7790 hCoV-19/USA/NY-NYUMC110/2020\|EPI_ISL_421722\|2020-0 | ···················· | ···················· | ···················· | ···················· | ···················· | ···················· | |
| A | 7791 hCoV-19/USA/NJ-NYUMC111/2020\|EPI_ISL_421723\|2020-0 | ···················· | ···················· | ···················· | ···················· | ···················· | ···················· | |
| A | 7792 hCoV-19/USA/NY-NYUMC112/2020\|EPI_ISL_421724\|2020-0 | ···················· | ···················· | ···················· | ···················· | ···················· | ···················· | |
| A | 7793 hCoV-19/USA/NY-NYUMC113/2020\|EPI_ISL_421725\|2020-0 | ···················· | ···················· | ···················· | ···················· | ···················· | ···················· | |
| A | 7794 hCoV-19/USA/NY-NYUMC114/2020\|EPI_ISL_421726\|2020-0 | ···················· | ···················· | ···················· | ···················· | ···················· | ···················· | |
| A | 7795 hCoV-19/USA/NY-NYUMC115/2020\|EPI_ISL_421727\|2020-0 | ···················· | ···················· | ···················· | ···················· | ···················· | ···················· | |
| A | 7796 hCoV-19/USA/NY-NYUMC116/2020\|EPI_ISL_421728\|2020-0 | ···················· | ···················· | ···················· | ···················· | ···················· | ···················· | |
| A | 7797 hCoV-19/USA/NY-NYUMC117/2020\|EPI_ISL_421729\|2020-0 | ···················· | ···················· | ···················· | ···················· | ···················· | ···················· | |
| A | 7798 hCoV-19/USA/NY-NYUMC118/2020\|EPI_ISL_421730\|2020-0 | ···················· | ···················· | ···················· | ···················· | ···················· | ···················· | |
| A | 7799 hCoV-19/USA/NY-NYUMC119/2020\|EPI_ISL_421731\|2020-0 | ···················· | ···················· | ···················· | ···················· | ···················· | ···················· | |
| A | 7800 hCoV-19/USA/NY-NYUMC120/2020\|EPI_ISL_421732\|2020-0 | ···················· | ···················· | ···················· | ···················· | ···················· | ···················· | |
| A | 7801 hCoV-19/Luxembourg/LNS7970956/2020\|EPI_ISL_421734\| | ···················· | ···················· | ···················· | ···················· | ···················· | ···················· | |
| A | 7802 hCoV-19/Spain/VC-FISABIO-77/2020\|EPI_ISL_425201\|20 | ···················· | ···················· | ···················· | ···················· | ···················· | ···················· | |
| A | 7803 hCoV-19/Spain/VC-FISABIO-79/2020\|EPI_ISL_425203\|20 | ···················· | ···················· | ···················· | ···················· | ···················· | ···················· | |
| A | 7804 hCoV-19/Spain/VC-FISABIO-81/2020\|EPI_ISL_425205\|20 | ···················· | ···················· | ···················· | ···················· | ···················· | ···················· | |
| A | 7805 hCoV-19/Spain/VC-FISABIO-86/2020\|EPI_ISL_425210\|20 | ···················· | ···················· | ···················· | ···················· | ···················· | ···················· | |
| A | 7806 hCoV-19/Spain/VC-FISABIO-88/2020\|EPI_ISL_425212\|20 | ···················· | ···················· | ···················· | ···················· | ···················· | ···················· | |
| A | 7807 hCoV-19/Spain/VC-FISABIO-89/2020\|EPI_ISL_425213\|20 | ···················· | ···················· | ···················· | ···················· | ···················· | ···················· | |
| A | 7808 hCoV-19/Spain/VC-FISABIO-90/2020\|EPI_ISL_425214\|20 | ···················· | ···················· | ···················· | ···················· | ···················· | ···················· | |
| A | 7809 hCoV-19/Spain/VC-FISABIO-92/2020\|EPI_ISL_425216\|20 | ···················· | ···················· | ···················· | ···················· | ···················· | ···················· | |
| A | 7810 hCoV-19/Spain/VC-FISABIO-93/2020\|EPI_ISL_425217\|20 | ···················· | ···················· | ···················· | ···················· | ···················· | ···················· | |
| A | 7811 hCoV-19/Spain/VC-FISABIO-94/2020\|EPI_ISL_425218\|20 | ···················· | ···················· | ···················· | ···················· | ···················· | ···················· | |
| A | 7812 hCoV-19/Spain/VC-FISABIO-95/2020\|EPI_ISL_425219\|20 | ···················· | ···················· | ···················· | ···················· | ···················· | ···················· | |
| A | 7813 hCoV-19/Spain/VC-FISABIO-96/2020\|EPI_ISL_425220\|20 | ···················· | ···················· | ···················· | ···················· | ···················· | ···················· | |
| A | 7814 hCoV-19/Spain/VC-FISABIO-97/2020\|EPI_ISL_425221\|20 | ···················· | ···················· | ···················· | ···················· | ···················· | ···················· | |
| A | 7815 hCoV-19/USA/NY-Wadsworth-10495-01/2020\|EPI_ISL_425 | ···················· | ···················· | ···················· | ···················· | ···················· | ···················· | |
| A | 7816 hCoV-19/Czech Republic/2556/2020\|EPI_ISL_425228\|20 | ···················· | ···················· | ···················· | ···················· | ···················· | ···················· | |
| A | 7817 hCoV-19/Czech Republic/2122/2020\|EPI_ISL_425229\|20 | ···················· | ···················· | ···················· | ···················· | ···················· | ···············A···· | |
| A | 7818 hCoV-19/England/CAMB-7374F/2020\|EPI_ISL_425231\|202 | ···················· | ···················A | ···················· | ···················· | ···················· | ···················· | 1 |
| A | 7819 hCoV-19/England/CAMB-7375E/2020\|EPI_ISL_425232\|202 | ···················· | ···················· | ···················· | ···················· | ···················· | ···················· | |

| | | | | | | | |
|---|---|---|---|---|---|---|---|
| A | 7820 hCoV-19/England/CAMB-7376D/2020\|EPI_ISL_425233\|202 | .................... | .................... | .................... | .................... | .................... | .................... |
| A | 7821 hCoV-19/England/CAMB-7377C/2020\|EPI_ISL_425234\|202 | .................... | .................... | .................... | .................... | .................... | .................... |
| A | 7822 hCoV-19/England/CAMB-7378B/2020\|EPI_ISL_425235\|202 | .................... | .................... | .................... | .................... | .................... | .................... |
| A | 7823 hCoV-19/England/CAMB-7379A/2020\|EPI_ISL_425236\|202 | .................... | .................... | .................... | .................... | .................... | .................... |
| A | 7825 hCoV-19/England/CAMB-737B8/2020\|EPI_ISL_425238\|202 | .................... | .................... | .................... | .................... | .................... | .................... |
| A | 7826 hCoV-19/England/CAMB-737C7/2020\|EPI_ISL_425239\|202 | .................... | .................... | .................... | .................... | .................... | .................... |
| A | 7827 hCoV-19/England/CAMB-737D6/2020\|EPI_ISL_425240\|202 | .................... | .................... | .................... | .................... | .................... | .................... |
| A | 7828 hCoV-19/England/CAMB-737F4/2020\|EPI_ISL_425242\|202 | .................... | .................... | .................... | .................... | .................... | .................... |
| A | 7829 hCoV-19/England/CAMB-73800/2020\|EPI_ISL_425243\|202 | .................... | .................... | .................... | .................... | .................... | .................... |
| A | 7830 hCoV-19/England/CAMB-7381F/2020\|EPI_ISL_425244\|202 | .................... | .................... | .................... | .................... | .................... | .................... |
| A | 7831 hCoV-19/England/CAMB-7386A/2020\|EPI_ISL_425246\|202 | .................... | .................... | .................... | .................... | .................... | .................... |
| A | 7832 hCoV-19/England/CAMB-73888/2020\|EPI_ISL_425248\|202 | .................... | .................... | .................... | .................... | .................... | .................... |
| A | 7833 hCoV-19/England/CAMB-738A6/2020\|EPI_ISL_425249\|202 | .................... | .................... | .................... | .................... | .................... | .................... |
| A | 7834 hCoV-19/England/CAMB-738B5/2020\|EPI_ISL_425250\|202 | .................... | .................... | .................... | .................... | .................... | .................... |
| A | 7835 hCoV-19/England/CAMB-738C4/2020\|EPI_ISL_425251\|202 | .................... | .................... | .................... | .................... | .................... | .................... |
| A | 7836 hCoV-19/England/CAMB-738D3/2020\|EPI_ISL_425252\|202 | .................... | .................... | .................... | .................... | .................... | .................... |
| A | 7837 hCoV-19/England/CAMB-738E2/2020\|EPI_ISL_425253\|202 | .................... | .................... | .................... | .................... | .................... | .................... |
| A | 7838 hCoV-19/England/CAMB-7391C/2020\|EPI_ISL_425255\|202 | .................... | .................... | .................... | .................... | .................... | .................... |
| A | 7839 hCoV-19/England/CAMB-73949/2020\|EPI_ISL_425257\|202 | .................... | .................... | .................... | .................... | .................... | .................... |
| A | 7840 hCoV-19/England/CAMB-73976/2020\|EPI_ISL_425258\|202 | .................... | .................... | .................... | .................... | .................... | .................... |
| A | 7841 hCoV-19/England/CAMB-73985/2020\|EPI_ISL_425259\|202 | .................... | .................... | .................... | .................... | .................... | .................... |
| A | 7842 hCoV-19/England/CAMB-73994/2020\|EPI_ISL_425260\|202 | .................... | .................... | .................... | .................... | .................... | .................... |
| A | 7843 hCoV-19/England/CAMB-739A3/2020\|EPI_ISL_425261\|202 | .................... | .................... | .................... | .................... | .................... | .................... |
| A | 7844 hCoV-19/England/CAMB-739B2/2020\|EPI_ISL_425262\|202 | .................... | .................... | .................... | .................... | .................... | .................... |
| A | 7845 hCoV-19/England/CAMB-739D0/2020\|EPI_ISL_425263\|202 | .................... | .................... | .................... | .................... | .................... | .................... |
| A | 7846 hCoV-19/England/CAMB-739EF/2020\|EPI_ISL_425264\|202 | .................... | .................... | .................... | .................... | .................... | .................... |
| A | 7847 hCoV-19/England/CAMB-73A0A/2020\|EPI_ISL_425266\|202 | .................... | .................... | .................... | .................... | .................... | .................... |
| A | 7848 hCoV-19/England/CAMB-73A19/2020\|EPI_ISL_425267\|202 | .................... | .................... | .................... | .................... | .................... | .................... |
| A | 7849 hCoV-19/England/CAMB-73A28/2020\|EPI_ISL_425268\|202 | .................... | .................... | .................... | .................... | .................... | .................... |
| A | 7850 hCoV-19/England/CAMB-73A37/2020\|EPI_ISL_425269\|202 | .................... | .................... | .................... | .................... | .................... | NN.................. |
| A | 7851 hCoV-19/USA/AZ-TG268099/2020\|EPI_ISL_426500\|2020-0 | .................... | .................... | .................... | .................... | .................... | .................... |
| A | 7852 hCoV-19/USA/AZ-TG268183/2020\|EPI_ISL_426501\|2020-0 | .................... | .................... | .................... | .................... | .................... | .................... |
| A | 7853 hCoV-19/USA/AZ-TG268188/2020\|EPI_ISL_426502\|2020-0 | .................... | .................... | .................... | .................... | .................... | .................... |
| A | 7854 hCoV-19/USA/AZ-TG268281/2020\|EPI_ISL_426503\|2020-0 | .................... | .................... | .................... | .................... | .................... | .................... |
| A | 7855 hCoV-19/USA/AZ-TG268282/2020\|EPI_ISL_426504\|2020-0 | .................... | .................... | .................... | .................... | .................... | .................... |
| A | 7856 hCoV-19/USA/AZ-TG268289/2020\|EPI_ISL_426505\|2020-0 | .................... | .................... | .................... | .................... | .................... | .................... |
| A | 7857 hCoV-19/USA/AZ-TG268350/2020\|EPI_ISL_426506\|2020-0 | .................... | .................... | .................... | .................... | .................... | .................... |
| A | 7858 hCoV-19/USA/AZ-TG268360/2020\|EPI_ISL_426507\|2020-0 | .................... | .................... | .................... | .................... | .................... | .................... |
| A | 7859 hCoV-19/USA/AZ-TG268417/2020\|EPI_ISL_426508\|2020-0 | .................... | .................... | .................... | .................... | .................... | .................... |
| A | 7860 hCoV-19/USA/AZ-TG268629/2020\|EPI_ISL_426509\|2020-0 | .................... | .................... | .................... | .................... | .................... | .................... |
| A | 7861 hCoV-19/USA/AZ-TG268657/2020\|EPI_ISL_426510\|2020-0 | .................... | .................... | .................... | .................... | .................... | .................... |
| A | 7862 hCoV-19/USA/AZ-TG268770/2020\|EPI_ISL_426511\|2020-0 | .................... | .................... | .................... | .................... | .................... | .................... |
| A | 7863 hCoV-19/USA/AZ-TG268893/2020\|EPI_ISL_426512\|2020-0 | .................... | .................... | .................... | .................... | .................... | .................... |
| A | 7864 hCoV-19/USA/AZ-TG268903/2020\|EPI_ISL_426513\|2020-0 | .................... | .................... | .................... | .................... | .................... | .................... |
| A | 7865 hCoV-19/USA/AZ-TG268905/2020\|EPI_ISL_426514\|2020-0 | .................... | .................... | .................... | .................... | .................... | .................... |
| A | 7866 hCoV-19/USA/AZ-TG268909/2020\|EPI_ISL_426515\|2020-0 | .................... | .................... | .................... | .................... | .................... | .................... |
| A | 7867 hCoV-19/USA/AZ-TG268911/2020\|EPI_ISL_426516\|2020-0 | .................... | .................... | .................... | .................... | .................... | .................... |
| A | 7868 hCoV-19/USA/AZ-TG268913/2020\|EPI_ISL_426517\|2020-0 | .................... | .................... | .................... | .................... | .................... | .................... |
| A | 7869 hCoV-19/USA/AZ-TG268915/2020\|EPI_ISL_426518\|2020-0 | .................... | .................... | .................... | .................... | .................... | .................... |
| A | 7870 hCoV-19/USA/AZ-TG268917/2020\|EPI_ISL_426519\|2020-0 | .................... | .................... | .................... | .................... | .................... | .................... |
| A | 7871 hCoV-19/USA/AZ-TG269027/2020\|EPI_ISL_426520\|2020-0 | .................... | .................... | .................... | .................... | .................... | .................... |
| A | 7872 hCoV-19/USA/AZ-TG269238/2020\|EPI_ISL_426521\|2020-0 | .................... | .................... | .................... | .................... | .................... | .................... |
| A | 7873 hCoV-19/USA/AZ-TG269290/2020\|EPI_ISL_426522\|2020-0 | .................... | .................... | .................... | .................... | .................... | .................... |
| A | 7874 hCoV-19/USA/AZ-TG269358/2020\|EPI_ISL_426523\|2020-0 | .................... | .................... | .................... | .................... | .................... | .................... |
| A | 7875 hCoV-19/USA/AZ-TG269439/2020\|EPI_ISL_426524\|2020-0 | .................... | .................... | .................... | .................... | .................... | .................... |
| A | 7876 hCoV-19/USA/AZ-TG269440/2020\|EPI_ISL_426525\|2020-0 | .................... | .................... | .................... | .................... | .................... | .................... |
| A | 7877 hCoV-19/USA/AZ-TG269824/2020\|EPI_ISL_426526\|2020-0 | .................... | .................... | .................... | .................... | .................... | .................... |
| A | 7878 hCoV-19/USA/AZ-TG269848/2020\|EPI_ISL_426527\|2020-0 | .................... | .................... | .................... | .................... | .................... | .................... |
| A | 7879 hCoV-19/USA/AZ-TG269856/2020\|EPI_ISL_426528\|2020-0 | .................... | .................... | .................... | .................... | .................... | .................... |
| A | 7880 hCoV-19/USA/AZ-TG269863/2020\|EPI_ISL_426529\|2020-0 | .................... | .................... | .................... | .................... | .................... | .................... |
| A | 7881 hCoV-19/USA/AZ-TG269875/2020\|EPI_ISL_426530\|2020-0 | .................... | .................... | .................... | .................... | .................... | .................... |
| A | 7882 hCoV-19/USA/AZ-TG269883/2020\|EPI_ISL_426531\|2020-0 | .................... | .................... | .................... | .................... | .................... | .................... |
| A | 7883 hCoV-19/USA/AZ-TG270152/2020\|EPI_ISL_426532\|2020-0 | .................... | .................... | .................... | .................... | .................... | .................... |
| A | 7884 hCoV-19/USA/AZ-TG270200/2020\|EPI_ISL_426533\|2020-0 | .................... | .................... | .................... | .................... | .................... | .................... |
| A | 7885 hCoV-19/USA/AZ-TG271435/2020\|EPI_ISL_426534\|2020-0 | .................... | .................... | .................... | .................... | .................... | .................... |
| A | 7886 hCoV-19/USA/AZ-TG271578/2020\|EPI_ISL_426535\|2020-0 | .................... | .................... | .................... | .................... | .................... | .................... |
| A | 7887 hCoV-19/USA/AZ-TG271591/2020\|EPI_ISL_426536\|2020-0 | .................... | .................... | .................... | .................... | .................... | .................... |
| A | 7888 hCoV-19/USA/AZ-TG271854/2020\|EPI_ISL_426537\|2020-0 | .................... | .................... | .................... | .................... | .................... | .................... |
| A | 7889 hCoV-19/USA/AZ-TG271856/2020\|EPI_ISL_426538\|2020-0 | .................... | .................... | .................... | .................... | .................... | .................... |
| A | 7890 hCoV-19/USA/AZ-TG271864/2020\|EPI_ISL_426540\|2020-0 | .................... | .................... | .................... | .................... | .................... | .................... |
| A | 7891 hCoV-19/USA/AZ-TG271870/2020\|EPI_ISL_426541\|2020-0 | .................... | .................... | .................... | .................... | .................... | .................... |
| A | 7892 hCoV-19/USA/AZ-TG271878/2020\|EPI_ISL_426542\|2020-0 | .................... | .................... | .................... | .................... | .................... | .................... |
| A | 7893 hCoV-19/USA/AZ-TG271880/2020\|EPI_ISL_426543\|2020-0 | .................... | .................... | .................... | .................... | .................... | .................... |
| A | 7894 hCoV-19/USA/AZ-TG271882/2020\|EPI_ISL_426544\|2020-0 | .................... | .................... | .................... | .................... | .................... | .................... |
| A | 7895 hCoV-19/USA/AZ-TG271884/2020\|EPI_ISL_426545\|2020-0 | .................... | .................... | .................... | .................... | .................... | .................... |
| A | 7896 hCoV-19/USA/AZ-TG271886/2020\|EPI_ISL_426546\|2020-0 | .................... | .................... | .................... | .................... | .................... | .................... |
| A | 7897 hCoV-19/USA/AZ-TG271888/2020\|EPI_ISL_426547\|2020-0 | .................... | .................... | .................... | .................... | .................... | .................... |
| A | 7898 hCoV-19/USA/AZ-TG271890/2020\|EPI_ISL_426548\|2020-0 | .................... | .................... | .................... | .................... | .................... | .................... |

| | | | | | | | | | |
|---|---|---|---|---|---|---|---|---|---|
| A | 7899 hCoV-19/USA/AZ-TG271892/2020\|EPI_ISL_426549\|2020-0 | ···················· | ···················· | ···················· | ···················· | ···················· | ···················· | | |
| A | 7900 hCoV-19/USA/AZ-TG271894/2020\|EPI_ISL_426550\|2020-0 | ···················· | ···················· | ···················· | ···················· | ···················· | ···················· | | |
| A | 7901 hCoV-19/USA/AZ-TG271896/2020\|EPI_ISL_426551\|2020-0 | ···················· | ···················· | ···················· | ···················· | ···················· | ···················· | | |
| A | 7902 hCoV-19/USA/AZ-TG271898/2020\|EPI_ISL_426552\|2020-0 | ···················· | ···················· | ···················· | ···················· | ···················· | ···················· | | |
| A | 7903 hCoV-19/USA/AZ-TG271900/2020\|EPI_ISL_426553\|2020-0 | ···················· | ···················· | ···················· | ···················· | ···················· | ···················· | | |
| A | 7904 hCoV-19/USA/AZ-TG271902/2020\|EPI_ISL_426554\|2020-0 | ···················· | ···················· | ···················· | ···················· | ···················· | ···················· | | |
| A | 7905 hCoV-19/USA/AZ-TG271922/2020\|EPI_ISL_426555\|2020-0 | ···················· | ···················· | ···················· | ···················· | ···················· | ···················· | | |
| A | 7906 hCoV-19/USA/AZ-TG272213/2020\|EPI_ISL_426556\|2020-0 | ···················· | ···················· | ···················· | ···················· | ···················· | ···················· | | |
| A | 7907 hCoV-19/USA/AZ-TG272215/2020\|EPI_ISL_426557\|2020-0 | ···················· | ···················· | ···················· | ···················· | ···················· | ···················· | | |
| A | 7908 hCoV-19/USA/AZ-TG273280/2020\|EPI_ISL_426558\|2020-0 | ···················· | ···················· | ···················· | ···················· | ···················· | ···················· | | |
| A | 7909 hCoV-19/USA/AZ-TG273282/2020\|EPI_ISL_426559\|2020-0 | ···················· | ···················· | ···················· | ···················· | ···················· | ···················· | | |
| A | 7910 hCoV-19/USA/AZ-TG273286/2020\|EPI_ISL_426560\|2020-0 | ···················· | ···················· | ···················· | ···················· | ···················· | ···················· | | |
| A | 7911 hCoV-19/USA/AZ-TG273294/2020\|EPI_ISL_426561\|2020-0 | ···················· | ···················· | ···················· | ···················· | ···················· | ···················· | | |
| A | 7912 hCoV-19/USA/AZ-TG273300/2020\|EPI_ISL_426562\|2020-0 | ···················· | ···················· | ···················· | ···················· | ···················· | ···················· | | |
| A | 7913 hCoV-19/USA/AZ-TG273302/2020\|EPI_ISL_426563\|2020-0 | ···················· | ···················· | ···················· | ···················· | ···················· | ···················· | | |
| A | 7914 hCoV-19/USA/AZ-TG273304/2020\|EPI_ISL_426564\|2020-0 | ···················· | ···················· | ···················· | ···················· | ···················· | ···················· | | |
| A | 7915 hCoV-19/USA/AZ-TG273308/2020\|EPI_ISL_426565\|2020-0 | ···················· | ···················· | ···················· | ···················· | ···················· | ···················· | | |
| A | 7916 hCoV-19/USA/AZ-TG273310/2020\|EPI_ISL_426566\|2020-0 | ···················· | ···················· | ···················· | ···················· | ···················· | ···················· | | |
| A | 7917 hCoV-19/USA/AZ-TG273314/2020\|EPI_ISL_426567\|2020-0 | ···················· | ···················· | ···················· | ···················· | ···················· | ···················· | | |
| A | 7918 hCoV-19/USA/AZ-TG273316/2020\|EPI_ISL_426568\|2020-0 | ···················· | ···················· | ···················· | ···················· | ···················· | ···················· | | |
| A | 7919 hCoV-19/USA/AZ-TG273318/2020\|EPI_ISL_426569\|2020-0 | ···················· | ···················· | ···················· | ···················· | ···················· | ···················· | | |
| A | 7920 hCoV-19/Brazil/DF-0001/2020\|EPI_ISL_426580\|2020-03 | ···················· | ···················· | ···················· | ···················· | ···················· | ···················· | | |
| A | 7921 hCoV-19/Czech Republic/IAB20-006-16/2020\|EPI_ISL_4 | ···················· | ···················· | ···················· | ···················· | ···················· | ···················· | | |
| A | 7922 hCoV-19/Australia/NSW87/2020\|EPI_ISL_427800\|2020-0 | ···················· | ···················· | ···················· | ···················· | ···················· | ···················· | | |
| A | 7923 hCoV-19/Australia/NSW173/2020\|EPI_ISL_427801\|2020- | ···················· | ···················· | ···················· | ···················· | ···················· | ···················· | | |
| A | 7924 hCoV-19/Australia/NSW141/2020\|EPI_ISL_427802\|2020- | ···················· | ···················· | ···················· | ···················· | ···················· | ···················· | | |
| A | 7925 hCoV-19/Australia/NSW186/2020\|EPI_ISL_427803\|2020- | ···················· | ···················· | ···················· | ···················· | ···················· | ···················· | | |
| A | 7926 hCoV-19/Australia/NSW108/2020\|EPI_ISL_427804\|2020- | ···················· | ···················· | ···················· | ···················· | ···················· | ···················· | | |
| A | 7927 hCoV-19/Germany/BY-ChVir-1020/2020\|EPI_ISL_450200\| | ···················· | ···················· | ···················· | ···················· | ···················· | ···················· | | |
| A | 7928 hCoV-19/Germany/BY-ChVir-1247/2020\|EPI_ISL_450201\| | ···················· | ···················· | ···················· | ···················· | ···················· | ···················· | | |
| A | 7929 hCoV-19/Germany/BY-ChVir-1248/2020\|EPI_ISL_450202\| | ···················· | ···················· | ···················· | ···················· | ···················· | ···················· | | |
| A | 7930 hCoV-19/Germany/BY-ChVir-1289/2020\|EPI_ISL_450203\| | ···················· | ···················· | ···················· | ···················· | ···················· | ···················· | | |
| A | 7931 hCoV-19/Germany/BY-ChVir-1356/2020\|EPI_ISL_450204\| | ···················· | ···················· | ···················· | ···················· | ···················· | ···················· | | |
| A | 7932 hCoV-19/Germany/BY-ChVir-1433/2020\|EPI_ISL_450205\| | ···················· | ···················· | ···················· | ···················· | ···················· | ···················· | | |
| A | 7933 hCoV-19/Germany/BY-ChVir-1459/2020\|EPI_ISL_450206\| | ···················· | ···················· | ···················· | ···················· | ···················· | ···················· | | |
| A | 7934 hCoV-19/Germany/BY-ChVir-1482/2020\|EPI_ISL_450207\| | ···················· | ···················· | ···················· | ···················· | ···················· | ···················· | | |
| A | 7935 hCoV-19/Germany/BY-ChVir-1483/2020\|EPI_ISL_450208\| | ···················· | ···················· | ···················· | ···················· | ···················· | ···················· | | |
| A | 7936 hCoV-19/Germany/BY-ChVir-1017/2020\|EPI_ISL_450209\| | ···················· | ···················· | ···················· | ···················· | ···················· | ···················· | | |
| A | 7937 hCoV-19/France/IDF-0515/2020\|EPI_ISL_408430\|2020-0 | ···················· | ···················· | ···················· | ···················· | ···················· | ···················· | | |
| A | 7938 hCoV-19/France/IDF-0626/2020\|EPI_ISL_408431\|2020-0 | ···················· | ···················· | ···················· | ···················· | ···················· | ···················· | | |
| A | 7939 hCoV-19/Chongqing/YC01/2020\|EPI_ISL_408478\|2020-01 | ···················· | ···················· | ···················· | ···················· | ···················· | ···················· | | |
| A | 7940 hCoV-19/Chongqing/ZX01/2020\|EPI_ISL_408479\|2020-01 | ···················· | ···················· | ···················· | ···················· | ···················· | ···················· | | |
| A | 7941 hCoV-19/Yunnan/IVDC-YN-003/2020\|EPI_ISL_408480\|202 | ···················· | ···················· | ···················· | ···················· | ···················· | ···················· | | |
| A | 7942 hCoV-19/Chongqing/IVDC-CQ-001/2020\|EPI_ISL_408481\| | ···················· | ···················· | ···················· | ···················· | ···················· | ···················· | | |
| A | 7943 hCoV-19/Shandong/IVDC-SD-001/2020\|EPI_ISL_408482\|2 | ···················· | ···················· | ···················· | ···················· | ···················· | ···················· | | |
| A | 7944 hCoV-19/Sichuan/IVDC-SC-001/2020\|EPI_ISL_408484\|20 | ···················· | ···················· | ···················· | ···················· | ···················· | ···················· | | |
| A | 7945 hCoV-19/Beijing/IVDC-BJ-005/2020\|EPI_ISL_408485\|20 | ···················· | ···················· | ···················· | ···················· | ···················· | ···················· | | |
| A | 7946 hCoV-19/Jiangxi/IVDC-JX-002/2020\|EPI_ISL_408486\|20 | ···················· | ···················· | ···················· | ···················· | ···················· | ···················· | | |
| A | 7947 hCoV-19/Jiangsu/IVDC-JS-001/2020\|EPI_ISL_408488\|20 | ···················· | ···················· | ···················· | ···················· | ···················· | ···················· | | |
| A | 7948 hCoV-19/Taiwan/NTU01/2020\|EPI_ISL_408489\|2020-01-3 | ···················· | ···················· | ···················· | ···················· | ···················· | ···················· | | |
| A | 7949 hCoV-19/Singapore/11/2020\|EPI_ISL_410719\|2020-02-0 | ···················· | ···················· | ···················· | ···················· | ···················· | ···················· | | |
| A | 7950 hCoV-19/France/IDF-0372-isl/2020\|EPI_ISL_410720\|20 | ···················· | ···················· | ···················· | ···················· | ···················· | ···················· | | |
| A | 7951 hCoV-19/pangolin/Guangdong/1/2019\|EPI_ISL_410721\|2 | ···················· | ···················· | ···················· | ···················· | ·TCTT·T·A··T········ | ···A··T··G·····A··A· | 1 | 1 |
| A | 7952 hCoV-19/Hong Kong/VM20002907/2020\|EPI_ISL_414517\|2 | ···················· | ···················· | ···················· | ···················· | ···················· | ···················· | | |
| A | 7953 hCoV-19/Hong Kong/VM20002868/2020\|EPI_ISL_414519\|2 | ···················· | ···················· | ···················· | ···················· | ···················· | ···················· | | |
| A | 7954 hCoV-19/Germany/BY-BIM-Pat2/2020\|EPI_ISL_414520\|20 | ···················· | ···················· | ···················· | ···················· | ···················· | ···················· | | |
| A | 7955 hCoV-19/Germany/BY-BIM-Pat3/2020\|EPI_ISL_414521\|20 | ···················· | ···················· | ···················· | ···················· | ···················· | ···················· | | |
| A | 7956 hCoV-19/England/200990002/2020\|EPI_ISL_414522\|2020 | ···················· | ···················· | ···················· | ···················· | ···················· | ···················· | | |
| A | 7957 hCoV-19/England/200990660/2020\|EPI_ISL_414523\|2020 | ···················· | ···················· | ···················· | ···················· | ···················· | ···················· | | |
| A | 7958 hCoV-19/England/200991076/2020\|EPI_ISL_414524\|2020 | ···T················ | ···················· | ···················· | ···················· | ···················· | ···················· | | |
| A | 7959 hCoV-19/England/201040081/2020\|EPI_ISL_414525\|2020 | ···················· | ···················· | ···················· | ···················· | ···················· | ···················· | | |
| A | 7960 hCoV-19/England/201040141/2020\|EPI_ISL_414526\|2020 | ···················· | ···················· | ···················· | ···················· | ···················· | ···················· | | |
| A | 7961 hCoV-19/Hong Kong/VM20002493-2/2020\|EPI_ISL_414527 | ···················· | ···················· | ···················· | ···················· | ···················· | ···················· | | |
| A | 7962 hCoV-19/Hong Kong/VM20002507-2/2020\|EPI_ISL_414528 | ···················· | ···················· | ···················· | ···················· | ···················· | ···················· | | |
| A | 7963 hCoV-19/Netherlands/NoordBrabant_20/2020\|EPI_ISL_4 | ···················· | ···················· | ···················· | ···················· | ···················· | ···················· | | |
| A | 7964 hCoV-19/Netherlands/NoordBrabant_21/2020\|EPI_ISL_4 | ···················· | ···················· | ···················· | ···················· | ···················· | ···················· | | |
| A | 7965 hCoV-19/Netherlands/NoordBrabant_22/2020\|EPI_ISL_4 | ···················· | ···················· | ···················· | ···················· | ···················· | ···················· | | |
| A | 7966 hCoV-19/Netherlands/NoordBrabant_23/2020\|EPI_ISL_4 | ···················· | ···················· | ···················· | ···················· | ···················· | ···················· | | |
| A | 7967 hCoV-19/Netherlands/NoordBrabant_25/2020\|EPI_ISL_4 | ···················· | ···················· | ···················· | ···················· | ···················· | ···················· | | |
| A | 7968 hCoV-19/Netherlands/NoordBrabant_26/2020\|EPI_ISL_4 | ···················· | ···················· | ···················· | ···················· | ···················· | ···················· | | |
| A | 7969 hCoV-19/Netherlands/NoordBrabant_27/2020\|EPI_ISL_4 | ···················· | ···················· | ···················· | ···················· | ···················· | ···················· | | |
| A | 7970 hCoV-19/USA/WA-UW63/2020\|EPI_ISL_415591\|2020-03-10 | ···················· | ···················· | ···················· | ···················· | ···················· | ···················· | | |
| A | 7971 hCoV-19/USA/WA-UW64/2020\|EPI_ISL_415592\|2020-03-09 | ···················· | ···················· | ···················· | ···················· | ···················· | ···················· | | |
| A | 7972 hCoV-19/USA/WA-UW66/2020\|EPI_ISL_415594\|2020-03-10 | ···················· | ···················· | ···················· | ···················· | ···················· | ···················· | | |
| A | 7973 hCoV-19/USA/WA-UW67/2020\|EPI_ISL_415595\|2020-03-09 | ···················· | ···················· | ···················· | ···················· | ···················· | ···················· | | |
| A | 7974 hCoV-19/USA/WA-UW68/2020\|EPI_ISL_415596\|2020-03-09 | ···················· | ···················· | ···················· | ···················· | ···················· | ···················· | | |
| A | 7975 hCoV-19/USA/WA-UW69/2020\|EPI_ISL_415597\|2020-03-10 | ···················· | ···················· | ···················· | ···················· | ···················· | ···················· | | |
| A | 7976 hCoV-19/USA/WA-UW70/2020\|EPI_ISL_415598\|2020-03-10 | ···················· | ···················· | ···················· | ···················· | ···················· | ···················· | | |

| | | | | | | | | | |
|---|---|---|---|---|---|---|---|---|---|
| A | 7977 | hCoV-19/USA/WA-UW71/2020\|EPI_ISL_415599\|2020-03-09 | .................... | .................... | .................... | .................... | .................... | .................... | |
| A | 7978 | hCoV-19/Singapore/23/2020\|EPI_ISL_420100\|2020-03-0 | .................... | .................... | .................... | .................... | .................... | .................... | |
| A | 7979 | hCoV-19/Singapore/24/2020\|EPI_ISL_420101\|2020-03-0 | .................... | .................... | .................... | .................... | .................... | .................... | |
| A | 7980 | hCoV-19/Singapore/25/2020\|EPI_ISL_420102\|2020-03-0 | .................... | .................... | .................... | .................... | .................... | .................... | |
| A | 7981 | hCoV-19/Singapore/26/2020\|EPI_ISL_420103\|2020-03-0 | .................... | .................... | .................... | .................... | .................... | .................... | |
| A | 7982 | hCoV-19/Singapore/27/2020\|EPI_ISL_420104\|2020-03-0 | .................... | .................... | .................... | .................... | .................... | .................... | |
| A | 7983 | hCoV-19/Singapore/28/2020\|EPI_ISL_420105\|2020-03-0 | .................... | .................... | .................... | .................... | .................... | .................... | |
| A | 7984 | hCoV-19/Singapore/29/2020\|EPI_ISL_420106\|2020-03-0 | .................... | .................... | .................... | .................... | .................... | .................... | |
| A | 7985 | hCoV-19/Singapore/30/2020\|EPI_ISL_420107\|2020-03-0 | .................... | .................... | .................... | .................... | .................... | .................... | |
| A | 7986 | hCoV-19/Singapore/31/2020\|EPI_ISL_420108\|2020-03-1 | .................... | .................... | .................... | .................... | .................... | .................... | |
| A | 7987 | hCoV-19/Singapore/32/2020\|EPI_ISL_420109\|2020-03-1 | .................... | .................... | .................... | .................... | .................... | .................... | |
| A | 7988 | hCoV-19/Singapore/33/2020\|EPI_ISL_420110\|2020-03-1 | .................... | .................... | .................... | .................... | .................... | .................... | |
| A | 7989 | hCoV-19/Singapore/34/2020\|EPI_ISL_420111\|2020-03-1 | .................... | .................... | .................... | .................... | .................... | .................... | |
| A | 7990 | hCoV-19/Spain/VC-FISABIO-27/2020\|EPI_ISL_420112\|20 | .................... | .................... | .................... | .................... | .................... | .................... | |
| A | 7991 | hCoV-19/Spain/VC-FISABIO-28/2020\|EPI_ISL_420113\|20 | .................... | .................... | .................... | .................... | .................... | .................... | |
| A | 7992 | hCoV-19/Spain/VC-FISABIO-29/2020\|EPI_ISL_420114\|20 | .................... | .................... | .................... | .................... | .................... | .................... | |
| A | 7993 | hCoV-19/Spain/VC-FISABIO-30/2020\|EPI_ISL_420115\|20 | .................... | .................... | .................... | .................... | .................... | .................... | |
| A | 7994 | hCoV-19/Spain/VC-FISABIO-31/2020\|EPI_ISL_420116\|20 | .................... | .................... | .................... | .................... | .................... | .................... | |
| A | 7995 | hCoV-19/Spain/VC-FISABIO-32/2020\|EPI_ISL_420117\|20 | .................... | .................... | .................... | .................... | .................... | .................... | |
| A | 7996 | hCoV-19/Spain/VC-FISABIO-33/2020\|EPI_ISL_420118\|20 | .................... | .................... | .................... | .................... | .................... | .................... | |
| A | 7997 | hCoV-19/Spain/VC-FISABIO-34/2020\|EPI_ISL_420119\|20 | .................... | .................... | .................... | .................... | .................... | .................... | |
| A | 7998 | hCoV-19/Spain/VC-FISABIO-35/2020\|EPI_ISL_420120\|20 | .................... | .................... | .................... | .................... | .................... | .................... | |
| A | 7999 | hCoV-19/Spain/VC-FISABIO-36/2020\|EPI_ISL_420121\|20 | .................... | .................... | .................... | .................... | .................... | .................... | |
| A | 8000 | hCoV-19/Spain/VC-FISABIO-37/2020\|EPI_ISL_420122\|20 | .................... | .................... | .................... | .................... | .................... | .................... | |
| A | 8001 | hCoV-19/Spain/VC-FISABIO-38/2020\|EPI_ISL_420123\|20 | .................... | .................... | .................... | .................... | .................... | .................... | |
| A | 8002 | hCoV-19/Spain/VC-FISABIO-39/2020\|EPI_ISL_420124\|20 | .................... | .................... | .................... | .................... | .................... | .................... | |
| A | 8003 | hCoV-19/Spain/VC-FISABIO-41/2020\|EPI_ISL_420125\|20 | .................... | .................... | .................... | .................... | .................... | .................... | |
| A | 8004 | hCoV-19/Spain/VC-FISABIO-42/2020\|EPI_ISL_420126\|20 | .................... | .................... | .................... | .................... | .................... | .................... | |
| A | 8005 | hCoV-19/Spain/VC-FISABIO-43/2020\|EPI_ISL_420127\|20 | .................... | .................... | .................... | .................... | .................... | .................... | |
| A | 8006 | hCoV-19/Spain/VC-FISABIO-44/2020\|EPI_ISL_420128\|20 | .................... | .................... | .................... | .................... | .................... | .................... | |
| A | 8007 | hCoV-19/Spain/VC-FISABIO-45/2020\|EPI_ISL_420129\|20 | .................... | .................... | .................... | .................... | .................... | .................... | |
| A | 8008 | hCoV-19/Spain/VC-FISABIO-49/2020\|EPI_ISL_420130\|20 | .................... | .................... | .................... | .................... | .................... | .................... | |
| A | 8009 | hCoV-19/Spain/VC-FISABIO-50/2020\|EPI_ISL_420131\|20 | .................... | .................... | .................... | .................... | .................... | .................... | |
| A | 8010 | hCoV-19/Spain/VC-FISABIO-51/2020\|EPI_ISL_420132\|20 | .................... | .................... | .................... | .................... | .................... | .................... | |
| A | 8011 | hCoV-19/Norway/1455/2020\|EPI_ISL_420135\|2020-02-28 | .................... | .................... | .................... | .................... | .................... | .................... | |
| A | 8012 | hCoV-19/USA/NY-PV08449/2020\|EPI_ISL_421400\|2020-03 | .................... | .................... | .................... | .................... | .................... | .................... | |
| A | 8013 | hCoV-19/USA/NY-PV08450/2020\|EPI_ISL_421401\|2020-03 | .................... | .................... | .................... | .................... | .................... | .................... | |
| A | 8014 | hCoV-19/USA/NY-PV08452/2020\|EPI_ISL_421402\|2020-03 | .................... | .................... | .................... | .................... | .................... | .................... | |
| A | 8015 | hCoV-19/USA/NY-PV08453/2020\|EPI_ISL_421403\|2020-03 | .................... | .................... | .................... | .................... | .................... | .................... | |
| A | 8016 | hCoV-19/USA/NY-PV08455/2020\|EPI_ISL_421404\|2020-03 | .................... | .................... | .................... | .................... | .................... | .................... | |
| A | 8017 | hCoV-19/USA/NY-PV08456/2020\|EPI_ISL_421405\|2020-03 | .................... | .................... | .................... | .................... | .................... | .................... | |
| A | 8018 | hCoV-19/USA/NY-PV08459/2020\|EPI_ISL_421406\|2020-03 | .................... | .................... | .................... | .................... | .................... | .................... | |
| A | 8019 | hCoV-19/USA/NY-PV08461/2020\|EPI_ISL_421407\|2020-03 | .................... | .................... | .................... | .................... | .................... | .................... | |
| A | 8020 | hCoV-19/USA/NY-PV08462/2020\|EPI_ISL_421408\|2020-03 | .................... | .................... | .................... | .................... | .................... | .................... | |
| A | 8021 | hCoV-19/USA/NY-PV08463/2020\|EPI_ISL_421409\|2020-03 | .................... | .................... | .................... | .................... | .................... | .................... | |
| A | 8022 | hCoV-19/USA/NY-PV08464/2020\|EPI_ISL_421410\|2020-03 | .................... | .................... | .................... | .................... | .................... | .................... | |
| A | 8023 | hCoV-19/USA/NY-PV08466/2020\|EPI_ISL_421411\|2020-03 | .................... | .................... | .................... | .................... | .................... | .................... | |
| A | 8024 | hCoV-19/USA/NY-PV08468/2020\|EPI_ISL_421412\|2020-03 | .................... | ......A............. | .................... | .................... | .................... | .................... | |
| A | 8025 | hCoV-19/USA/NY-PV08469/2020\|EPI_ISL_421413\|2020-03 | .................... | .................... | .................... | .................... | .................... | .................... | |
| A | 8026 | hCoV-19/USA/NY-PV08471/2020\|EPI_ISL_421414\|2020-03 | .................... | .................... | .................... | .................... | .................... | .................... | |
| A | 8027 | hCoV-19/USA/NY-PV08472/2020\|EPI_ISL_421415\|2020-03 | .................... | .................... | .................... | .................... | .................... | .................... | |
| A | 8028 | hCoV-19/USA/NY-PV08473/2020\|EPI_ISL_421416\|2020-03 | .................... | .................... | .................... | .................... | .................... | .................... | |
| A | 8029 | hCoV-19/USA/NY-PV08474/2020\|EPI_ISL_421417\|2020-03 | .................... | .................... | .................... | .................... | .................... | .................... | |
| A | 8030 | hCoV-19/USA/NY-PV08477/2020\|EPI_ISL_421419\|2020-03 | .................... | .................... | .................... | .................... | .................... | .................... | |
| A | 8031 | hCoV-19/USA/NY-PV08478/2020\|EPI_ISL_421420\|2020-03 | .................... | ...................A | .................... | .................... | .................... | .................... | 1 |
| A | 8032 | hCoV-19/USA/NY-PV08481/2020\|EPI_ISL_421421\|2020-03 | .................... | .................... | .................... | .................... | .................... | .................... | |
| A | 8033 | hCoV-19/USA/NY-PV08485/2020\|EPI_ISL_421422\|2020-03 | .................... | .................... | .................... | .................... | .................... | .................... | |
| A | 8034 | hCoV-19/USA/NY-PV08486/2020\|EPI_ISL_421423\|2020-03 | .................... | .................... | .................... | .................... | .................... | .................... | |
| A | 8035 | hCoV-19/USA/NY-PV08487/2020\|EPI_ISL_421424\|2020-03 | .................... | .................... | .................... | .................... | .................... | .................... | |
| A | 8036 | hCoV-19/USA/NY-PV08489/2020\|EPI_ISL_421425\|2020-03 | .................... | .................... | .................... | .................... | .................... | .................... | |
| A | 8037 | hCoV-19/USA/NY-PV08490/2020\|EPI_ISL_421426\|2020-03 | .................... | .................... | .................... | .................... | .................... | .................... | |
| A | 8038 | hCoV-19/USA/NY-PV08491/2020\|EPI_ISL_421427\|2020-03 | .................... | .................... | .................... | .................... | .................... | .................... | |
| A | 8039 | hCoV-19/USA/NY-PV08492/2020\|EPI_ISL_421428\|2020-03 | .................... | .................... | .................... | .................... | .................... | .................... | |
| A | 8040 | hCoV-19/USA/NY-PV08493/2020\|EPI_ISL_421429\|2020-03 | .................... | .................... | .................... | .................... | .................... | .................... | |
| A | 8041 | hCoV-19/USA/NY-PV08494/2020\|EPI_ISL_421430\|2020-03 | .................... | .................... | .................... | .................... | .................... | .................... | |
| A | 8042 | hCoV-19/USA/NY-PV08495/2020\|EPI_ISL_421431\|2020-03 | .................... | .................... | .................... | .................... | .................... | .................... | |
| A | 8043 | hCoV-19/USA/NY-PV08496/2020\|EPI_ISL_421432\|2020-03 | .................... | .................... | .................... | .................... | .................... | .................... | |
| A | 8044 | hCoV-19/USA/NY-PV08498/2020\|EPI_ISL_421433\|2020-03 | .................... | .................... | .................... | .................... | .................... | .................... | |
| A | 8045 | hCoV-19/USA/NY-PV08500/2020\|EPI_ISL_421434\|2020-03 | .................... | .................... | .................... | .................... | .................... | .................... | |
| A | 8046 | hCoV-19/USA/NY-PV09000/2020\|EPI_ISL_421435\|2020-03 | .................... | .................... | .................... | .................... | .................... | .................... | |
| A | 8047 | hCoV-19/Portugal/PT0043/2020\|EPI_ISL_421446\|2020-0 | .................... | .................... | .................... | .................... | .................... | .................... | |
| A | 8048 | hCoV-19/Portugal/PT0044/2020\|EPI_ISL_421447\|2020-0 | .................... | .................... | .................... | .................... | .................... | .................... | |
| A | 8049 | hCoV-19/Portugal/PT0045/2020\|EPI_ISL_421448\|2020-0 | .................... | .................... | .................... | .................... | .................... | .................... | |
| A | 8050 | hCoV-19/Portugal/PT0046/2020\|EPI_ISL_421449\|2020-0 | .................... | .................... | .................... | .................... | .................... | .................... | |
| A | 8051 | hCoV-19/Portugal/PT0047/2020\|EPI_ISL_421450\|2020-0 | .................... | .................... | .................... | .................... | .................... | .................... | |
| A | 8052 | hCoV-19/Portugal/PT0048/2020\|EPI_ISL_421451\|2020-0 | .................... | .................... | .................... | .................... | .................... | .................... | |
| A | 8053 | hCoV-19/Portugal/PT0049/2020\|EPI_ISL_421452\|2020-0 | .................... | .................... | .................... | .................... | .................... | .................... | |
| A | 8054 | hCoV-19/Portugal/PT0050/2020\|EPI_ISL_421453\|2020-0 | .................... | .................... | .................... | .................... | .................... | .................... | |

| | | | | | | | | |
|---|---|---|---|---|---|---|---|---|
| A | 8055 | hCoV-19/Portugal/PT0051/2020\|EPI_ISL_421454\|2020-0 | ···················· | ···················· | ···················· | ···················· | ···················· | ···················· |
| A | 8056 | hCoV-19/Portugal/PT0052/2020\|EPI_ISL_421455\|2020-0 | ···················· | ···················· | ···················· | ···················· | ···················· | ···················· |
| A | 8057 | hCoV-19/Portugal/PT0053/2020\|EPI_ISL_421456\|2020-0 | ···················· | ···················· | ···················· | ···················· | ···················· | ···················· |
| A | 8058 | hCoV-19/Portugal/PT0054/2020\|EPI_ISL_421457\|2020-0 | ···················· | ···················· | ···················· | ···················· | ···················· | ···················· |
| A | 8059 | hCoV-19/Portugal/PT0055/2020\|EPI_ISL_421458\|2020-0 | ···················· | ···················· | ···················· | ···················· | ···················· | ···················· |
| A | 8060 | hCoV-19/Portugal/PT0056/2020\|EPI_ISL_421459\|2020-0 | ···················· | ···················· | ···················· | ···················· | ···················· | ···················· |
| A | 8061 | hCoV-19/Portugal/PT0057/2020\|EPI_ISL_421460\|2020-0 | ···················· | ·A·················· | ···················· | ···················· | ···················· | ···················· |
| A | 8062 | hCoV-19/Portugal/PT0058/2020\|EPI_ISL_421461\|2020-0 | ···················· | ···················· | ···················· | ···················· | ···················· | ···················· |
| A | 8063 | hCoV-19/Portugal/PT0059/2020\|EPI_ISL_421462\|2020-0 | ···················· | ···················· | ···················· | ···················· | ···················· | ···················· |
| A | 8064 | hCoV-19/Portugal/PT0060/2020\|EPI_ISL_421463\|2020-0 | ···················· | ···················· | ···················· | ···················· | ···················· | ···················· |
| A | 8065 | hCoV-19/Portugal/PT0061/2020\|EPI_ISL_421464\|2020-0 | ···················· | ···················· | ···················· | ···················· | ···················· | ···················· |
| A | 8066 | hCoV-19/Portugal/PT0062/2020\|EPI_ISL_421465\|2020-0 | ···················· | ···················· | ···················· | ···················· | ···················· | ···················· |
| A | 8067 | hCoV-19/Portugal/PT0063/2020\|EPI_ISL_421466\|2020-0 | ···················· | ···················· | ···················· | ···················· | ···················· | ···················· |
| A | 8068 | hCoV-19/Netherlands/NA_165/2020\|EPI_ISL_422700\|202 | ···················· | ···················· | ···················· | ···················· | ···················· | ···················· |
| A | 8069 | hCoV-19/Netherlands/NA_166/2020\|EPI_ISL_422701\|202 | ···················· | ···················· | ···················· | ···················· | ···················· | ···················· |
| A | 8070 | hCoV-19/Netherlands/NA_167/2020\|EPI_ISL_422702\|202 | ···················· | ···················· | ···················· | ···················· | ···················· | ···················· |
| A | 8071 | hCoV-19/Netherlands/NA_168/2020\|EPI_ISL_422703\|202 | ···················· | ···················· | ···················· | ···················· | ···················· | ···················· |
| A | 8072 | hCoV-19/Netherlands/NA_169/2020\|EPI_ISL_422704\|202 | ···················· | ···················· | ···················· | ···················· | ····T··············· | ···················· |
| A | 8073 | hCoV-19/Netherlands/NA_170/2020\|EPI_ISL_422705\|202 | ···················· | ···················· | ···················· | ···················· | ····T··············· | ···················· |
| A | 8074 | hCoV-19/Netherlands/NA_171/2020\|EPI_ISL_422706\|202 | ···················· | ···················· | ···················· | ···················· | ···················· | ···················· |
| A | 8075 | hCoV-19/Netherlands/NA_172/2020\|EPI_ISL_422707\|202 | ···················· | ···················· | ···················· | ···················· | ···················· | ···················· |
| A | 8076 | hCoV-19/Netherlands/NA_173/2020\|EPI_ISL_422708\|202 | ···················· | ···················· | ···················· | ···················· | ···················· | ···················· |
| A | 8077 | hCoV-19/Netherlands/NA_174/2020\|EPI_ISL_422709\|202 | ···················· | ···················· | ···················· | ···················· | ···················· | ···················· |
| A | 8078 | hCoV-19/Netherlands/NA_176/2020\|EPI_ISL_422711\|202 | ···················· | ···················· | ···················· | ···················· | ···················· | ···················· |
| A | 8079 | hCoV-19/Netherlands/NA_177/2020\|EPI_ISL_422712\|202 | ···················· | ···················· | ···················· | ···················· | ···················· | ···················· |
| A | 8080 | hCoV-19/Netherlands/NA_179/2020\|EPI_ISL_422714\|202 | ···················· | ···················· | ···················· | ···················· | ···················· | ···················· |
| A | 8081 | hCoV-19/Netherlands/NA_180/2020\|EPI_ISL_422715\|202 | ···················· | ···················· | ···················· | ···················· | ···················· | ···················· |
| A | 8082 | hCoV-19/Netherlands/NA_181/2020\|EPI_ISL_422716\|202 | ···················· | ···················· | ···················· | ···················· | ···················· | ···················· |
| A | 8083 | hCoV-19/Netherlands/NA_182/2020\|EPI_ISL_422717\|202 | ···················· | ···················· | ···················· | ···················· | ···················· | ···················· |
| A | 8084 | hCoV-19/Netherlands/NA_183/2020\|EPI_ISL_422718\|202 | ···················· | ···················· | ···················· | ···················· | ···················· | ···················· |
| A | 8085 | hCoV-19/Netherlands/NA_184/2020\|EPI_ISL_422719\|202 | ···················· | ···················· | ···················· | ···················· | ···················· | ···················· |
| A | 8086 | hCoV-19/Netherlands/NA_185/2020\|EPI_ISL_422720\|202 | ···················· | ···················· | ···················· | ···················· | ···················· | ···················· |
| A | 8087 | hCoV-19/Netherlands/NA_186/2020\|EPI_ISL_422721\|202 | ···················· | ···················· | ···················· | ···················· | ···················· | ···················· |
| A | 8088 | hCoV-19/Netherlands/NA_187/2020\|EPI_ISL_422722\|202 | ···················· | ···················· | ···················· | ···················· | ···················· | ···················· |
| A | 8089 | hCoV-19/Netherlands/NA_189/2020\|EPI_ISL_422724\|202 | ···················· | ···················· | ···················· | ···················· | ···················· | ···················· |
| A | 8090 | hCoV-19/Netherlands/NA_190/2020\|EPI_ISL_422725\|202 | ···················· | ···················· | ···················· | ···················· | ···················· | ···················· |
| A | 8091 | hCoV-19/Netherlands/NA_191/2020\|EPI_ISL_422726\|202 | ···················· | ···················· | ···················· | ···················· | ···················· | ···················· |
| A | 8092 | hCoV-19/Netherlands/NA_192/2020\|EPI_ISL_422727\|202 | ···················· | ···················· | ···················· | ···················· | ···················· | ···················· |
| A | 8093 | hCoV-19/Netherlands/NA_193/2020\|EPI_ISL_422728\|202 | ···················· | ···················· | ···················· | ···················· | ···················· | ···················· |
| A | 8094 | hCoV-19/Netherlands/NA_194/2020\|EPI_ISL_422729\|202 | ···················· | ···················· | ···················· | ···················· | ···················· | ···················· |
| A | 8095 | hCoV-19/Netherlands/NA_195/2020\|EPI_ISL_422730\|202 | ···················· | ···················· | ···················· | ···················· | ···················· | ···················· |
| A | 8096 | hCoV-19/Netherlands/NA_197/2020\|EPI_ISL_422732\|202 | ···················· | ···················· | ···················· | ···················· | ···················· | ···················· |
| A | 8097 | hCoV-19/Netherlands/NA_198/2020\|EPI_ISL_422733\|202 | ···················· | ···················· | ···················· | ···················· | ···················· | ···················· |
| A | 8098 | hCoV-19/Netherlands/NA_199/2020\|EPI_ISL_422734\|202 | ···················· | ···················· | ···················· | ···················· | ···················· | ···················· |
| A | 8099 | hCoV-19/Netherlands/NA_200/2020\|EPI_ISL_422735\|202 | ···················· | ···················· | ···················· | ···················· | ···················· | ···················· |
| A | 8100 | hCoV-19/Netherlands/NA_201/2020\|EPI_ISL_422736\|202 | ···················· | ···················· | ···················· | ···················· | ···················· | ···················· |
| A | 8101 | hCoV-19/Netherlands/NA_202/2020\|EPI_ISL_422737\|202 | ···················· | ···················· | ···················· | ···················· | ···················· | ···················· |
| A | 8102 | hCoV-19/Netherlands/NA_203/2020\|EPI_ISL_422738\|202 | ···················· | ···················· | ···················· | ···················· | ···················· | ···················· |
| A | 8103 | hCoV-19/Netherlands/NA_206/2020\|EPI_ISL_422741\|202 | ···················· | ···················· | ···················· | ···················· | ···················· | ···················· |
| A | 8104 | hCoV-19/Netherlands/NA_207/2020\|EPI_ISL_422742\|202 | ···················· | ···················· | ···················· | ···················· | ···················· | ···················· |
| A | 8105 | hCoV-19/Netherlands/NA_209/2020\|EPI_ISL_422744\|202 | ···················· | ···················· | ···················· | ···················· | ···················· | ···················· |
| A | 8106 | hCoV-19/Netherlands/NA_210/2020\|EPI_ISL_422745\|202 | ···················· | ···················· | ···················· | ···················· | ···················· | ···················· |
| A | 8107 | hCoV-19/Netherlands/NA_211/2020\|EPI_ISL_422746\|202 | ···················· | ···················· | ···················· | ···················· | ···················· | ···················· |
| A | 8108 | hCoV-19/Netherlands/NA_212/2020\|EPI_ISL_422747\|202 | ···················· | ···················· | ···················· | ···················· | ···················· | ···················· |
| A | 8109 | hCoV-19/Netherlands/NA_213/2020\|EPI_ISL_422748\|202 | ···················· | ···················· | ···················· | ···················· | ···················· | ···················· |
| A | 8110 | hCoV-19/Netherlands/NA_214/2020\|EPI_ISL_422749\|202 | ···················· | ···················· | ···················· | ···················· | ···················· | ···················· |
| A | 8111 | hCoV-19/Netherlands/NA_215/2020\|EPI_ISL_422750\|202 | ···················· | ···················· | ···················· | ···················· | ···················· | ···················· |
| A | 8112 | hCoV-19/Netherlands/NA_216/2020\|EPI_ISL_422751\|202 | ···················· | ···················· | ···················· | ···················· | ···················· | ···················· |
| A | 8113 | hCoV-19/Netherlands/NA_217/2020\|EPI_ISL_422752\|202 | ···················· | ···················· | ···················· | ···················· | ···················· | ···················· |
| A | 8114 | hCoV-19/Netherlands/NA_218/2020\|EPI_ISL_422753\|202 | ···················· | ·A·················· | ···················· | ···················· | ···················· | ···················· |
| A | 8115 | hCoV-19/Netherlands/NA_219/2020\|EPI_ISL_422754\|202 | ···················· | ···················· | ···················· | ···················· | ···················· | ···················· |
| A | 8116 | hCoV-19/Netherlands/NA_220/2020\|EPI_ISL_422755\|202 | ···················· | ···················· | ···················· | ···················· | ···················· | ···················· |
| A | 8117 | hCoV-19/Netherlands/NA_221/2020\|EPI_ISL_422756\|202 | ···················· | ···················· | ···················· | ···················· | ···················· | ···················· |
| A | 8118 | hCoV-19/Netherlands/NA_223/2020\|EPI_ISL_422758\|202 | ···················· | ···················· | ···················· | ···················· | ···················· | ···················· |
| A | 8119 | hCoV-19/Netherlands/NA_224/2020\|EPI_ISL_422759\|202 | ···················· | ···················· | ···················· | ···················· | ···················· | ···················· |
| A | 8120 | hCoV-19/Netherlands/NA_225/2020\|EPI_ISL_422760\|202 | ···················· | ···················· | ···················· | ···················· | ···················· | ···················· |
| A | 8121 | hCoV-19/Netherlands/NA_226/2020\|EPI_ISL_422761\|202 | ···················· | ···················· | ···················· | ···················· | ···················· | ···················· |
| A | 8122 | hCoV-19/Netherlands/NA_227/2020\|EPI_ISL_422762\|202 | ···················· | ···················· | ···················· | ···················· | ···················· | ···················· |
| A | 8123 | hCoV-19/Netherlands/NA_229/2020\|EPI_ISL_422764\|202 | ···················· | ···················· | ···················· | ···················· | ···················· | ···················· |
| A | 8124 | hCoV-19/Netherlands/NA_230/2020\|EPI_ISL_422765\|202 | ···················· | ···················· | ···················· | ···················· | ···················· | ···················· |
| A | 8125 | hCoV-19/Netherlands/NA_231/2020\|EPI_ISL_422766\|202 | ···················· | ···················· | ···················· | ···················· | ···················· | ···················· |
| A | 8126 | hCoV-19/Netherlands/NA_233/2020\|EPI_ISL_422767\|202 | ···················· | ···················· | ···················· | ···················· | ···················· | ···················· |
| A | 8127 | hCoV-19/Netherlands/NA_234/2020\|EPI_ISL_422768\|202 | ···················· | ···················· | ···················· | ···················· | ···················· | ···················· |
| A | 8128 | hCoV-19/Netherlands/NA_235/2020\|EPI_ISL_422769\|202 | ···················· | ···················· | ···················· | ···················· | ···················· | ···················· |
| A | 8129 | hCoV-19/Netherlands/NA_236/2020\|EPI_ISL_422770\|202 | ···················· | ···················· | ···················· | ···················· | ···················· | ···················· |
| A | 8130 | hCoV-19/Netherlands/NA_237/2020\|EPI_ISL_422771\|202 | ···················· | ···················· | ···················· | ···················· | ···················· | ···················· |
| A | 8131 | hCoV-19/Netherlands/NA_238/2020\|EPI_ISL_422772\|202 | ···················· | ···················· | ···················· | ···················· | ···················· | ···················· |
| A | 8132 | hCoV-19/Netherlands/NA_240/2020\|EPI_ISL_422774\|202 | ···················· | ···················· | ···················· | ···················· | ···················· | ···················· |

| | | | | | | | | |
|---|---|---|---|---|---|---|---|---|
| A | 8133 | hCoV-19/Netherlands/NA_242/2020\|EPI_ISL_422776\|202 | .................... | .................... | .................... | .................... | .................... | .................... |
| A | 8134 | hCoV-19/Netherlands/NA_243/2020\|EPI_ISL_422777\|202 | .................... | .................... | .................... | .................... | .................... | .................... |
| A | 8135 | hCoV-19/Netherlands/NA_244/2020\|EPI_ISL_422778\|202 | .................... | .................... | .................... | .................... | .................... | .................... |
| A | 8136 | hCoV-19/Netherlands/NA_250/2020\|EPI_ISL_422784\|202 | .................... | .................... | .................... | .................... | .................... | .................... |
| A | 8137 | hCoV-19/Netherlands/NA_251/2020\|EPI_ISL_422785\|202 | .................... | .................... | .................... | .................... | .................... | .................... |
| A | 8138 | hCoV-19/Netherlands/NA_253/2020\|EPI_ISL_422787\|202 | .................... | .................... | .................... | .................... | .................... | .................... |
| A | 8139 | hCoV-19/Netherlands/NA_254/2020\|EPI_ISL_422788\|202 | .................... | .................... | .................... | .................... | .................... | .................... |
| A | 8140 | hCoV-19/Netherlands/NA_256/2020\|EPI_ISL_422790\|202 | .................... | .................... | .................... | .................... | .................... | .................... |
| A | 8141 | hCoV-19/Netherlands/NA_257/2020\|EPI_ISL_422791\|202 | .................... | .................... | .................... | .................... | .................... | .................... |
| A | 8142 | hCoV-19/Netherlands/NA_258/2020\|EPI_ISL_422792\|202 | .................... | .................... | .................... | .................... | .................... | .................... |
| A | 8143 | hCoV-19/Netherlands/NA_259/2020\|EPI_ISL_422793\|202 | .................... | .................... | .................... | .................... | .................... | .................... |
| A | 8144 | hCoV-19/Netherlands/NA_260/2020\|EPI_ISL_422794\|202 | .................... | .................... | .................... | .................... | .................... | .................... |
| A | 8145 | hCoV-19/Netherlands/NA_261/2020\|EPI_ISL_422795\|202 | .................... | .................... | .................... | .................... | .................... | .................... |
| A | 8146 | hCoV-19/Netherlands/NA_262/2020\|EPI_ISL_422796\|202 | .................... | .A.................. | .................... | .................... | .................... | .................... |
| A | 8147 | hCoV-19/Netherlands/NA_36/2020\|EPI_ISL_422797\|2020 | .................... | .................... | .................... | .................... | .................... | .................... |
| A | 8148 | hCoV-19/Latvia/05/2020\|EPI_ISL_426285\|2020-03-30 | .................... | .................... | .................... | .................... | .................... | .................... |
| A | 8149 | hCoV-19/Latvia/06/2020\|EPI_ISL_426286\|2020-03-30 | .................... | .................... | .................... | .................... | .................... | .................... |
| A | 8150 | hCoV-19/Latvia/07/2020\|EPI_ISL_426287\|2020-03-30 | .................... | .................... | .................... | .................... | .................... | .................... |
| A | 8151 | hCoV-19/Latvia/08/2020\|EPI_ISL_426288\|2020-03-30 | .................... | .................... | .................... | .................... | .................... | .................... |
| A | 8152 | hCoV-19/Latvia/09/2020\|EPI_ISL_426289\|2020-03-30 | .................... | .................... | .................... | .................... | .................... | .................... |
| A | 8153 | hCoV-19/USA/NY-WCMP10E06P/2020\|EPI_ISL_427570\|2020 | .................... | .................... | .................... | .................... | .................... | .................... |
| A | 8154 | hCoV-19/USA/NY-WCMP10E07P/2020\|EPI_ISL_427571\|2020 | .................... | .................... | .................... | .................... | .................... | .................... |
| A | 8155 | hCoV-19/USA/NY-WCMP10F04P/2020\|EPI_ISL_427572\|2020 | .................... | .................... | .................... | .................... | .................... | .................... |
| A | 8156 | hCoV-19/USA/NY-WCMP10F06P/2020\|EPI_ISL_427573\|2020 | .................... | .................... | .................... | .................... | .................... | .................... |
| A | 8157 | hCoV-19/USA/NY-WCMP10F09P/2020\|EPI_ISL_427574\|2020 | .................... | .................... | .................... | .................... | .................... | .................... |
| A | 8158 | hCoV-19/USA/NY-WCMP10F10P/2020\|EPI_ISL_427575\|2020 | .................... | .................... | .................... | .................... | .................... | .................... |
| A | 8159 | hCoV-19/USA/NY-WCMP10G05P/2020\|EPI_ISL_427576\|2020 | .................... | .................... | .................... | .................... | .................... | .................... |
| A | 8160 | hCoV-19/USA/NY-WCMP10G06P/2020\|EPI_ISL_427577\|2020 | .................... | .................... | .................... | .................... | .................... | .................... |
| A | 8161 | hCoV-19/USA/NY-WCMP10G10P/2020\|EPI_ISL_427578\|2020 | .................... | .................... | .................... | .................... | .................... | .................... |
| A | 8162 | hCoV-19/USA/NY-WCMP10G11P/2020\|EPI_ISL_427579\|2020 | .................... | .................... | .................... | .................... | .................... | .................... |
| A | 8163 | hCoV-19/USA/NY-WCMP10H01P/2020\|EPI_ISL_427580\|2020 | .................... | .................... | .................... | .................... | .................... | .................... |
| A | 8164 | hCoV-19/USA/NY-WCMP10H04P/2020\|EPI_ISL_427581\|2020 | .................... | .................... | .................... | .................... | .................... | .................... |
| A | 8165 | hCoV-19/USA/NY-WCMP10H08P/2020\|EPI_ISL_427582\|2020 | .................... | .................... | .................... | .................... | .................... | .................... |
| A | 8166 | hCoV-19/USA/NY-WCMP11A03P/2020\|EPI_ISL_427583\|2020 | .................... | .................... | .................... | .................... | .................... | .................... |
| A | 8167 | hCoV-19/USA/NY-WCMP11A06P/2020\|EPI_ISL_427584\|2020 | .................... | .................... | .................... | .................... | .................... | .................... |
| A | 8168 | hCoV-19/USA/NY-WCMP11B03P/2020\|EPI_ISL_427585\|2020 | .................... | .................... | .................... | .................... | .................... | .................... |
| A | 8169 | hCoV-19/USA/NY-WCMP11B04P/2020\|EPI_ISL_427586\|2020 | .................... | .................... | .................... | .................... | .................... | .................... |
| A | 8170 | hCoV-19/USA/NY-WCMP11B07P/2020\|EPI_ISL_427587\|2020 | .................... | .................... | .................... | .................... | .................... | .................... |
| A | 8171 | hCoV-19/USA/NY-WCMP11C03P/2020\|EPI_ISL_427588\|2020 | .................... | .................... | .................... | .................... | .................... | .................... |
| A | 8172 | hCoV-19/USA/NY-WCMP11C04P/2020\|EPI_ISL_427589\|2020 | .................... | .................... | .................... | .................... | .................... | .................... |
| A | 8173 | hCoV-19/USA/NY-WCMP11C05P/2020\|EPI_ISL_427590\|2020 | .................... | .................... | .................... | .................... | .................... | .................... |
| A | 8174 | hCoV-19/USA/NY-WCMP11D01N/2020\|EPI_ISL_427591\|2020 | .................... | .................... | .................... | .................... | .................... | .................... |
| A | 8175 | hCoV-19/USA/NY-WCMP11E04P/2020\|EPI_ISL_427592\|2020 | .................... | .................... | .................... | .................... | .................... | .................... |
| A | 8176 | hCoV-19/USA/NY-WCMP11E05P/2020\|EPI_ISL_427593\|2020 | .................... | .................... | .................... | .................... | .................... | .................... |
| A | 8177 | hCoV-19/USA/NY-WCMP11E09P/2020\|EPI_ISL_427594\|2020 | .................... | .................... | .................... | .................... | .................... | .................... |
| A | 8178 | hCoV-19/USA/NY-WCMP11F04P/2020\|EPI_ISL_427595\|2020 | .................... | .................... | .................... | .................... | .................... | .................... |
| A | 8179 | hCoV-19/USA/NY-WCMP11F06P/2020\|EPI_ISL_427596\|2020 | .................... | .................... | .................... | .................... | .................... | .................... |
| A | 8180 | hCoV-19/USA/NY-WCMP11F07P/2020\|EPI_ISL_427597\|2020 | .................... | .................... | .................... | .................... | .................... | .................... |
| A | 8181 | hCoV-19/USA/NY-WCMP11H06P/2020\|EPI_ISL_427598\|2020 | .................... | .................... | .................... | .................... | .................... | .................... |
| A | 8182 | hCoV-19/USA/NY-WCMP12B04P/2020\|EPI_ISL_427599\|2020 | .................... | .................... | .................... | .................... | .................... | .................... |
| A | 8183 | hCoV-19/Australia/NSW82/2020\|EPI_ISL_427805\|2020-0 | .................... | .................... | .................... | .................... | .................... | .................... |
| A | 8184 | hCoV-19/Australia/NSW196/2020\|EPI_ISL_427806\|2020- | .................... | .................... | .................... | .................... | .................... | .................... |
| A | 8185 | hCoV-19/Australia/NSW207/2020\|EPI_ISL_427807\|2020- | .................... | .................... | .................... | .................... | .................... | .................... |
| A | 8186 | hCoV-19/Australia/NSW127/2020\|EPI_ISL_427808\|2020- | .................... | .................... | .................... | .................... | .................... | .................... |
| A | 8187 | hCoV-19/South Korea/KCDC2015/2020\|EPI_ISL_427809\|2 | .................... | .................... | .................... | .................... | .................... | .................... |
| A | 8188 | hCoV-19/South Korea/KCDC2016/2020\|EPI_ISL_427810\|2 | .................... | .................... | .................... | .................... | .................... | .................... |
| A | 8189 | hCoV-19/South Korea/KCDC2017/2020\|EPI_ISL_427811\|2 | .................... | .................... | .................... | .................... | .................... | .................... |
| A | 8190 | hCoV-19/South Korea/KCDC2018/2020\|EPI_ISL_427812\|2 | .................... | .................... | .................... | .................... | .................... | .................... |
| A | 8191 | hCoV-19/South Korea/KCDC2019/2020\|EPI_ISL_427813\|2 | .................... | .................... | .................... | .................... | .................... | .................... |
| A | 8192 | hCoV-19/Russia/StPetersburg-RII4706S/2020\|EPI_ISL_ | .................... | .................... | .................... | .................... | .................... | .................... |
| A | 8193 | hCoV-19/USA/WA1/2020\|EPI_ISL_404895\|2020-01-19 | .................... | .................... | .................... | .................... | .................... | .................... |
| A | 8194 | hCoV-19/Singapore/7/2020\|EPI_ISL_410713\|2020-01-27 | .................... | ......A............. | .................... | .................... | .................... | .................... |
| A | 8195 | hCoV-19/Singapore/8/2020\|EPI_ISL_410714\|2020-02-03 | .................... | .................... | .................... | .................... | .................... | .................... |
| A | 8196 | hCoV-19/Singapore/9/2020\|EPI_ISL_410715\|2020-02-04 | .................... | .................... | .................... | .................... | .................... | .................... |
| A | 8197 | hCoV-19/Singapore/10/2020\|EPI_ISL_410716\|2020-02-0 | .................... | .................... | .................... | .................... | .................... | .................... |
| A | 8198 | hCoV-19/Australia/QLD03/2020\|EPI_ISL_410717\|2020-0 | .................... | .................... | .................... | .................... | .................... | .................... |
| A | 8199 | hCoV-19/Australia/QLD04/2020\|EPI_ISL_410718\|2020-0 | .................... | .................... | .................... | .................... | .................... | .................... |
| A | 8200 | hCoV-19/Netherlands/NoordBrabant_48/2020\|EPI_ISL_4 | .................... | .................... | .................... | .................... | .................... | .................... |
| A | 8201 | hCoV-19/Netherlands/NoordBrabant_49/2020\|EPI_ISL_4 | .................... | .................... | .................... | .................... | .................... | .................... |
| A | 8202 | hCoV-19/Netherlands/NoordBrabant_51/2020\|EPI_ISL_4 | .................... | .................... | .................... | .................... | .................... | .................... |
| A | 8203 | hCoV-19/Netherlands/NoordBrabant_52/2020\|EPI_ISL_4 | .................... | .................... | .................... | .................... | .................... | .................... |
| A | 8204 | hCoV-19/Netherlands/NoordBrabant_53/2020\|EPI_ISL_4 | .................... | .................... | .................... | .................... | .................... | .................... |
| A | 8205 | hCoV-19/Netherlands/NoordBrabant_54/2020\|EPI_ISL_4 | .................... | .................... | .................... | .................... | .................... | .................... |
| A | 8206 | hCoV-19/Netherlands/NoordBrabant_55/2020\|EPI_ISL_4 | .................... | .................... | .................... | .................... | .................... | .................... |
| A | 8207 | hCoV-19/Netherlands/NoordBrabant_56/2020\|EPI_ISL_4 | .................... | .................... | .................... | .................... | .................... | .................... |
| A | 8208 | hCoV-19/Netherlands/NoordBrabant_57/2020\|EPI_ISL_4 | .................... | .................... | .................... | .................... | .................... | .................... |
| A | 8209 | hCoV-19/Netherlands/NoordBrabant_58/2020\|EPI_ISL_4 | .................... | .................... | .................... | .................... | .................... | .................... |
| A | 8210 | hCoV-19/Netherlands/NoordBrabant_59/2020\|EPI_ISL_4 | .................... | .................... | .................... | .................... | .................... | .................... |

| | | | | | | | | | |
|---|---|---|---|---|---|---|---|---|---|
| A | 8211 hCoV-19/Netherlands/NoordBrabant_61/2020\|EPI_ISL_4 | ···················· | ···················· | ····················· | ···················· | ···················· | ···················· | | |
| A | 8212 hCoV-19/Netherlands/NoordBrabant_62/2020\|EPI_ISL_4 | ···················· | ···················· | ····················· | ···················· | ···················· | ···················· | | |
| A | 8213 hCoV-19/Netherlands/NoordBrabant_63/2020\|EPI_ISL_4 | ···················· | ···················· | ····················· | ···················· | ···················· | ···················· | | |
| A | 8214 hCoV-19/Netherlands/NoordBrabant_64/2020\|EPI_ISL_4 | ···················· | ···················· | ····················· | ···················· | ···················· | ···················· | | |
| A | 8215 hCoV-19/Netherlands/NoordBrabant_65/2020\|EPI_ISL_4 | ···················· | ···················· | ····················· | ···················· | ···················· | ···················· | | |
| A | 8216 hCoV-19/Netherlands/NoordBrabant_66/2020\|EPI_ISL_4 | ···················· | ···················· | ····················· | ···················· | ···················· | ···················· | | |
| A | 8217 hCoV-19/Netherlands/NoordBrabant_67/2020\|EPI_ISL_4 | ···················· | ···················· | ····················· | ···················· | ···················· | ···················· | | |
| A | 8218 hCoV-19/Netherlands/NoordBrabant_68/2020\|EPI_ISL_4 | ···················· | ···················· | ····················· | ···················· | ···················· | ···················· | | |
| A | 8219 hCoV-19/Netherlands/NoordHolland_3/2020\|EPI_ISL_41 | ···················· | ···················· | ····················· | ···················· | ···················· | ···················· | | |
| A | 8220 hCoV-19/Netherlands/Utrecht_17/2020\|EPI_ISL_415526 | ···················· | ···················· | ····················· | ···················· | ···················· | ···················· | | |
| A | 8221 hCoV-19/Netherlands/Utrecht_18/2020\|EPI_ISL_415527 | ···················· | ···················· | ····················· | ···················· | ···················· | ···················· | | |
| A | 8222 hCoV-19/Netherlands/ZuidHolland_25/2020\|EPI_ISL_41 | ···················· | ···················· | ····················· | ···················· | ···················· | ···················· | | |
| A | 8223 hCoV-19/Netherlands/ZuidHolland_26/2020\|EPI_ISL_41 | ···················· | ···················· | ····················· | ···················· | ···················· | ···················· | | |
| A | 8224 hCoV-19/Netherlands/ZuidHolland_27/2020\|EPI_ISL_41 | ···················· | ···················· | ····················· | ···················· | ···················· | ···················· | | |
| A | 8225 hCoV-19/Netherlands/ZuidHolland_28/2020\|EPI_ISL_41 | ···················· | ···················· | ····················· | ···················· | ···················· | ···················· | | |
| A | 8226 hCoV-19/Netherlands/ZuidHolland_29/2020\|EPI_ISL_41 | ···················· | ···················· | ····················· | ···················· | ···················· | ···················· | | |
| A | 8227 hCoV-19/Netherlands/ZuidHolland_30/2020\|EPI_ISL_41 | ···················· | ···················· | ····················· | ···················· | ···················· | ···················· | | |
| A | 8228 hCoV-19/Netherlands/ZuidHolland_31/2020\|EPI_ISL_41 | ···················· | ···················· | ····················· | ···················· | ···················· | ···················· | | |
| A | 8229 hCoV-19/USA/UT-01/2020\|EPI_ISL_415539\|2020-03-13 | ···················· | ···················· | ····················· | ···················· | ···················· | ···················· | | |
| A | 8230 hCoV-19/USA/UT-03/2020\|EPI_ISL_415541\|2020-03-13 | ···················· | ···················· | ····················· | ···················· | ···················· | ···················· | | |
| A | 8231 hCoV-19/USA/UT-04/2020\|EPI_ISL_415542\|2020-03-13 | ···················· | ···················· | ····················· | ···················· | ···················· | ···················· | | |
| A | 8232 hCoV-19/USA/UT-05/2020\|EPI_ISL_415543\|2020-03-13 | ···················· | ···················· | ····················· | ···················· | ···················· | ···················· | | |
| A | 8233 hCoV-19/USA/UT-06/2020\|EPI_ISL_415544\|2020-03-13 | ···················· | ···················· | ····················· | ···················· | ···················· | ···················· | | |
| A | 8234 hCoV-19/Canada/BC_13297/2020\|EPI_ISL_415578\|2020-0 | ···················· | ···················· | ····················· | ···················· | ···················· | ···················· | | |
| A | 8235 hCoV-19/Canada/BC_17397/2020\|EPI_ISL_415580\|2020-0 | ···················· | ···················· | ····················· | ···················· | ···················· | ···················· | | |
| A | 8236 hCoV-19/Canada/BC_02421/2020\|EPI_ISL_415581\|2020-0 | ···················· | ···················· | ····················· | ···················· | ···················· | ···················· | | |
| A | 8237 hCoV-19/Canada/BC_35720/2020\|EPI_ISL_415582\|2020-0 | ···················· | ···················· | ····················· | ···················· | ···················· | ···················· | | |
| A | 8238 hCoV-19/Canada/BC_40860/2020\|EPI_ISL_415583\|2020-0 | ···················· | ···················· | ····················· | ···················· | ···················· | ···················· | | |
| A | 8239 hCoV-19/Canada/BC_41851/2020\|EPI_ISL_415584\|2020-0 | ···················· | ···················· | ····················· | ···················· | ···················· | ···················· | | |
| A | 8242 hCoV-19/Canada/BC_83109/2020\|EPI_ISL_415588\|2020-0 | ···················· | ···················· | ····················· | ···················· | ···············N···N | NN·N················ | 1 | 1 |
| A | 8244 hCoV-19/Malaysia/MKAK-CL-2020-5047/2020\|EPI_ISL_41 | ···················· | ···················· | ····················· | ···················· | ···················· | ···················· | | |
| A | 8245 hCoV-19/Malaysia/MKAK-CL-2020-5096/2020\|EPI_ISL_41 | ···················· | ···················· | ····················· | ···················· | ···················· | ···················· | | |
| A | 8246 hCoV-19/Malaysia/MKAK-CL-2020-6430/2020\|EPI_ISL_41 | ···················· | ···················· | ····················· | ···················· | ···················· | ···················· | | |
| A | 8247 hCoV-19/USA/VA-DCLS-0033/2020\|EPI_ISL_420019\|2020- | ···················· | ···················· | ····················· | ···················· | ···················· | ···················· | | |
| A | 8248 hCoV-19/USA/VA-DCLS-0036/2020\|EPI_ISL_420020\|2020- | ···················· | ···················· | ····················· | ···················· | ···················· | ···················· | | |
| A | 8249 hCoV-19/USA/VA-DCLS-0039/2020\|EPI_ISL_420021\|2020- | ···················· | ···················· | ····················· | ···················· | ···················· | ···················· | | |
| A | 8250 hCoV-19/USA/VA-DCLS-0040/2020\|EPI_ISL_420022\|2020- | ···················· | ···················· | ····················· | ···················· | ···················· | ···················· | | |
| A | 8251 hCoV-19/USA/VA-DCLS-0042/2020\|EPI_ISL_420023\|2020- | ···················· | ···················· | ····················· | ···················· | ···················· | ···················· | | |
| A | 8252 hCoV-19/USA/VA-DCLS-0043/2020\|EPI_ISL_420024\|2020- | ···················· | ···················· | ····················· | ···················· | ···················· | ···················· | | |
| A | 8253 hCoV-19/USA/VA-DCLS-0044/2020\|EPI_ISL_420025\|2020- | ···················· | ···················· | ····················· | ···················· | ···················· | ···················· | | |
| A | 8254 hCoV-19/USA/VA-DCLS-0045/2020\|EPI_ISL_420026\|2020- | ···················· | ···················· | ····················· | ···················· | ···················· | ···················· | | |
| A | 8255 hCoV-19/USA/VA-DCLS-0046/2020\|EPI_ISL_420027\|2020- | ···················· | ···················· | ····················· | ···················· | ···················· | ···················· | | |
| A | 8256 hCoV-19/USA/VA-DCLS-0018/2020\|EPI_ISL_420028\|2020- | ···················· | ···················· | ····················· | ···················· | ···················· | ···················· | | |
| A | 8257 hCoV-19/USA/VA-DCLS-0019/2020\|EPI_ISL_420029\|2020- | ···················· | ···················· | ····················· | ···················· | ···················· | ···················· | | |
| A | 8258 hCoV-19/DRC/214/2020\|EPI_ISL_420030\|2020-03-21 | ···················· | ···················· | ····················· | ···················· | ···················· | ···················· | | |
| A | 8259 hCoV-19/DRC/215/2020\|EPI_ISL_420031\|2020-03-21 | ···················· | ···················· | ····················· | ···················· | ···················· | ···················· | | |
| A | 8260 hCoV-19/DRC/236/2020\|EPI_ISL_420032\|2020-03-22 | ···················· | ···················· | ····················· | ···················· | ···················· | ···················· | | |
| A | 8261 hCoV-19/DRC/241/2020\|EPI_ISL_420033\|2020-03-22 | ···················· | ···················· | ····················· | ···················· | ···················· | ···················· | | |
| A | 8262 hCoV-19/DRC/243/2020\|EPI_ISL_420034\|2020-03-22 | ···················· | ···················· | ····················· | ···················· | ···················· | ···················· | | |
| A | 8263 hCoV-19/DRC/248/2020\|EPI_ISL_420035\|2020-03-22 | ···················· | ···················· | ····················· | ···················· | ···················· | ···················· | | |
| A | 8264 hCoV-19/Australia/VIC256/2020\|EPI_ISL_420036\|2020- | ···················· | ···················· | ····················· | ···················· | ···················· | ···················· | | |
| A | 8265 hCoV-19/USA/NY-PV08121/2020\|EPI_ISL_421350\|2020-03 | ···················· | ···················· | ····················· | ···················· | ···················· | ···················· | | |
| A | 8266 hCoV-19/USA/NY-PV08122/2020\|EPI_ISL_421351\|2020-03 | ···················· | ···················· | ····················· | ···················· | ···················· | ···················· | | |
| A | 8267 hCoV-19/USA/NY-PV08123/2020\|EPI_ISL_421352\|2020-03 | ···················· | ···················· | ····················· | ···················· | ···················· | ···················· | | |
| A | 8268 hCoV-19/USA/NY-PV08124/2020\|EPI_ISL_421353\|2020-03 | ···················· | ···················· | ····················· | ···················· | ···················· | ···················· | | |
| A | 8269 hCoV-19/USA/NY-PV08125/2020\|EPI_ISL_421354\|2020-03 | ···················· | ···················· | ····················· | ···················· | ···················· | ···················· | | |
| A | 8270 hCoV-19/USA/NY-PV08127/2020\|EPI_ISL_421355\|2020-03 | ···················· | ···················· | ····················· | ···················· | ···················· | ···················· | | |
| A | 8271 hCoV-19/USA/NY-PV08135/2020\|EPI_ISL_421357\|2020-03 | ···················· | ·············A······ | ····················· | ···················· | ···················· | ···················· | | |
| A | 8272 hCoV-19/USA/NY-PV08136/2020\|EPI_ISL_421358\|2020-03 | ···················· | ·············A······ | ····················· | ···················· | ···················· | ···················· | | |
| A | 8273 hCoV-19/USA/NY-PV08137/2020\|EPI_ISL_421359\|2020-03 | ···················· | ·············A······ | ····················· | ···················· | ···················· | ···················· | | |
| A | 8274 hCoV-19/USA/NY-PV08138/2020\|EPI_ISL_421360\|2020-03 | ···················· | ···················· | ····················· | ···················· | ···················· | ···················· | | |
| A | 8275 hCoV-19/USA/NY-PV08143/2020\|EPI_ISL_421363\|2020-03 | ···················· | ···················· | ····················· | ···················· | ···················· | ···················· | | |
| A | 8276 hCoV-19/USA/NY-PV08146/2020\|EPI_ISL_421364\|2020-03 | ···················· | ···················· | ····················· | ···················· | ···················· | ···················· | | |
| A | 8277 hCoV-19/USA/NY-PV08148/2020\|EPI_ISL_421365\|2020-03 | ···················· | ···················· | ····················· | ···················· | ···················· | ···················· | | |
| A | 8278 hCoV-19/USA/NY-PV08149/2020\|EPI_ISL_421366\|2020-03 | ···················· | ···················· | ····················· | ···················· | ···················· | ···················· | | |
| A | 8279 hCoV-19/USA/NY-PV08150/2020\|EPI_ISL_421367\|2020-03 | ···················· | ···················· | ····················· | ···················· | ···················· | ···················· | | |
| A | 8280 hCoV-19/Netherlands/NA_136/2020\|EPI_ISL_422680\|202 | ···················· | ···················· | ····················· | ···················· | ···················· | ···················· | | |
| A | 8281 hCoV-19/Netherlands/NA_137/2020\|EPI_ISL_422681\|202 | ···················· | ···················· | ····················· | ···················· | ···················· | ···················· | | |
| A | 8282 hCoV-19/Netherlands/NA_138/2020\|EPI_ISL_422682\|202 | ···················· | ···················· | ····················· | ···················· | ···················· | ···················· | | |
| A | 8283 hCoV-19/Netherlands/NA_144/2020\|EPI_ISL_422684\|202 | ···················· | ···················· | ····················· | ···················· | ···················· | ···················· | | |
| A | 8284 hCoV-19/Netherlands/NA_145/2020\|EPI_ISL_422685\|202 | ···················· | ·············A······ | ····················· | ···················· | ···················· | ···················· | | |
| A | 8285 hCoV-19/Netherlands/NA_150/2020\|EPI_ISL_422686\|202 | ···················· | ···················· | ····················· | ···················· | ···················· | ···················· | | |
| A | 8286 hCoV-19/Netherlands/NA_152/2020\|EPI_ISL_422687\|202 | ···················· | ···················· | ····················· | ···················· | ···················· | ···················· | | |
| A | 8287 hCoV-19/Netherlands/NA_153/2020\|EPI_ISL_422688\|202 | ···················· | ···················· | ····················· | ···················· | ···················· | ···················· | | |
| A | 8288 hCoV-19/Netherlands/NA_154/2020\|EPI_ISL_422689\|202 | ···················· | ···················· | ····················· | ···················· | ···················· | ···················· | | |
| A | 8289 hCoV-19/Netherlands/NA_155/2020\|EPI_ISL_422690\|202 | ···················· | ···················· | ····················· | ···················· | ···················· | ···················· | | |
| A | 8290 hCoV-19/Netherlands/NA_156/2020\|EPI_ISL_422691\|202 | ···················· | ···················· | ····················· | ···················· | ···················· | ···················· | | |
| A | 8291 hCoV-19/Netherlands/NA_157/2020\|EPI_ISL_422692\|202 | ···················· | ···················· | ····················· | ···················· | ···················· | ···················· | | |

A 8292 hCoV-19/Netherlands/NA_158/2020|EPI_ISL_422693|202 .................... .................... .................... .................... .................... ....................
A 8293 hCoV-19/Netherlands/NA_159/2020|EPI_ISL_422694|202 .................... .................... .................... .................... .................... ....................
A 8294 hCoV-19/Netherlands/NA_160/2020|EPI_ISL_422695|202 .................... .................... .................... .................... .................... ....................
A 8295 hCoV-19/Netherlands/NA_161/2020|EPI_ISL_422696|202 .................... .................... .................... .................... .................... ....................
A 8296 hCoV-19/Netherlands/NA_162/2020|EPI_ISL_422697|202 .................... .................... .................... .................... .................... ....................
A 8297 hCoV-19/Netherlands/NA_163/2020|EPI_ISL_422698|202 .................... .................... .................... .................... .................... ....................
A 8298 hCoV-19/USA/NY-Wadsworth-11735-01/2020|EPI_ISL_426 .................... .................... .................... .................... .................... ....................
A 8299 hCoV-19/USA/NY-Wadsworth-11717-01/2020|EPI_ISL_426 .................... .................... .................... .................... .................... ....................
A 8300 hCoV-19/USA/NY-Wadsworth-11207-01/2020|EPI_ISL_426 .................... .................... .................... .................... .................... ....................
A 8301 hCoV-19/USA/NY-Wadsworth-12883-02/2020|EPI_ISL_426 .................... .................... .................... .................... .................... ....................
A 8302 hCoV-19/USA/NY-Wadsworth-13468-02/2020|EPI_ISL_426 .................... .................... .................... .................... .................... ....................
A 8303 hCoV-19/USA/NY-Wadsworth-27073-01/2020|EPI_ISL_426 .................... .................... .................... .................... .................... ....................
A 8304 hCoV-19/USA/NY-Wadsworth-16080-01/2020|EPI_ISL_426 .................... .................... .................... .................... .................... ....................
A 8305 hCoV-19/USA/NY-Wadsworth-27804-01/2020|EPI_ISL_426 .................... .................... .................... .................... .................... ....................
A 8306 hCoV-19/USA/NY-Wadsworth-31488-01/2020|EPI_ISL_426 .................... .................... .................... .................... .................... ....................
A 8307 hCoV-19/USA/NY-Wadsworth-32851-01/2020|EPI_ISL_426 .................... .................... .................... .................... .................... ....................
A 8308 hCoV-19/USA/NY-NYUMC207/2020|EPI_ISL_427502|2020-0 .................... .................... .................... .................... .................... ....................
A 8309 hCoV-19/USA/NY-NYUMC208/2020|EPI_ISL_427503|2020-0 .................... .................... .................... .................... .................... ....................
A 8310 hCoV-19/USA/NY-NYUMC209/2020|EPI_ISL_427504|2020-0 .................... .................... .................... .................... .................... ....................
A 8311 hCoV-19/USA/NY-NYUMC210/2020|EPI_ISL_427505|2020-0 .................... .................... .................... .................... .................... ....................
A 8312 hCoV-19/USA/NY-NYUMC212/2020|EPI_ISL_427507|2020-0 .................... .................... .................... .................... .................... ....................
A 8313 hCoV-19/USA/NY-NYUMC213/2020|EPI_ISL_427508|2020-0 .................... .................... .................... .................... .................... ....................
A 8314 hCoV-19/USA/NY-NYUMC214/2020|EPI_ISL_427509|2020-0 .................... .................... .................... .................... .................... ....................
A 8315 hCoV-19/USA/NY-NYUMC215/2020|EPI_ISL_427510|2020-0 .................... .................... .................... .................... .................... ....................
A 8316 hCoV-19/USA/NY-NYUMC216/2020|EPI_ISL_427511|2020-0 .................... .................... .................... .................... .................... ....................
A 8317 hCoV-19/USA/NY-NYUMC217/2020|EPI_ISL_427512|2020-0 .................... .................... .................... .................... .................... ....................
A 8318 hCoV-19/USA/NY-NYUMC218/2020|EPI_ISL_427513|2020-0 .................... .................... .................... .................... .................... ....................
A 8319 hCoV-19/USA/NY-NYUMC219/2020|EPI_ISL_427514|2020-0 .................... .................... .................... .................... .................... ....................
A 8320 hCoV-19/USA/NY-NYUMC220/2020|EPI_ISL_427515|2020-0 .................... .................... .................... .................... .................... ....................
A 8321 hCoV-19/USA/NY-NYUMC221/2020|EPI_ISL_427516|2020-0 .................... .................... .................... .................... .................... ....................
A 8322 hCoV-19/USA/NY-NYUMC223/2020|EPI_ISL_427518|2020-0 .................... .................... .................... .................... .................... ....................
A 8323 hCoV-19/USA/NY-NYUMC224/2020|EPI_ISL_427519|2020-0 .................... .................... .................... .................... .................... ....................
A 8324 hCoV-19/USA/NY-NYUMC225/2020|EPI_ISL_427520|2020-0 .................... .................... .................... .................... .................... ....................
A 8325 hCoV-19/USA/NY-NYUMC226/2020|EPI_ISL_427521|2020-0 .................... .................... .................... .................... .................... ....................
A 8326 hCoV-19/USA/NY-NYUMC176/2020|EPI_ISL_427524|2020-0 .................... .................... .................... .................... .................... ....................
A 8327 hCoV-19/USA/NY-WCMP2C03P/2020|EPI_ISL_427526|2020- .................... .................... .................... .................... .................... ....................
A 8328 hCoV-19/USA/NY-WCMP2D02P/2020|EPI_ISL_427527|2020- .................... .................... .................... .................... .................... ....................
A 8329 hCoV-19/USA/NY-WCMP2D04P/2020|EPI_ISL_427528|2020- .................... .................... .................... .................... .................... ....................
A 8330 hCoV-19/USA/NY-WCMP2H03P/2020|EPI_ISL_427529|2020- .................... .................... .................... .................... .................... ....................
A 8331 hCoV-19/USA/NY-WCMP3E02P/2020|EPI_ISL_427530|2020- .................... .................... .................... .................... .................... ....................
A 8332 hCoV-19/USA/NY-WCMP4A02N/2020|EPI_ISL_427531|2020- .................... .................... .................... .................... .................... ....................
A 8333 hCoV-19/USA/NY-WCMP4B06P/2020|EPI_ISL_427532|2020- .................... .................... .................... .................... .................... ....................
A 8334 hCoV-19/USA/NY-WCMP4C01P/2020|EPI_ISL_427533|2020- .................... .................... .................... .................... .................... ....................
A 8335 hCoV-19/USA/NY-WCMP4C02N/2020|EPI_ISL_427534|2020- .................... .................... .................... .................... .................... ....................
A 8336 hCoV-19/USA/NY-WCMP4D01P/2020|EPI_ISL_427535|2020- .................... .................... .................... .................... .................... ....................
A 8337 hCoV-19/USA/NY-WCMP4D04P/2020|EPI_ISL_427536|2020- .................... .................... .................... .................... .................... ....................
A 8338 hCoV-19/USA/NY-WCMP4E05P/2020|EPI_ISL_427537|2020- .................... .................... .................... .................... .................... ....................
A 8339 hCoV-19/USA/NY-WCMP4G04P/2020|EPI_ISL_427538|2020- .................... .................... .................... .................... .................... ....................
A 8340 hCoV-19/USA/NY-WCMP4H03P/2020|EPI_ISL_427539|2020- .................... .................... .................... .................... .................... ....................
A 8341 hCoV-19/USA/NY-WCMP5F01P/2020|EPI_ISL_427540|2020- .................... .................... .................... .................... .................... ....................
A 8342 hCoV-19/USA/NY-WCMP5G03P/2020|EPI_ISL_427541|2020- .................... .................... .................... .................... .................... ....................
A 8343 hCoV-19/USA/NY-WCMP5H02P/2020|EPI_ISL_427542|2020- .................... .................... .................... .................... .................... ....................
A 8344 hCoV-19/USA/NY-WCMP5H03P/2020|EPI_ISL_427543|2020- .................... .................... .................... .................... .................... ....................
A 8345 hCoV-19/USA/NY-WCMP9A05P/2020|EPI_ISL_427544|2020- .................... .................... .................... .................... .................... ....................
A 8346 hCoV-19/USA/NY-WCMP9A08P/2020|EPI_ISL_427545|2020- .................... .................... .................... .................... .................... ....................
A 8347 hCoV-19/USA/NY-WCMP9B05P/2020|EPI_ISL_427546|2020- .................... .................... .................... .................... .................... ....................
A 8348 hCoV-19/USA/NY-WCMP9B06P/2020|EPI_ISL_427547|2020- .................... .................... .................... .................... .................... ....................
A 8349 hCoV-19/USA/NY-WCMP9B08P/2020|EPI_ISL_427548|2020- .................... .................... .................... .................... .................... ....................
A 8350 hCoV-19/USA/NY-WCMP9B09P/2020|EPI_ISL_427549|2020- .................... .................... .................... .................... .................... ....................
A 8351 hCoV-19/USA/NY-WCMP9D03P/2020|EPI_ISL_427550|2020- .................... .................... .................... .................... .................... ....................
A 8352 hCoV-19/USA/NY-WCMP9E04P/2020|EPI_ISL_427551|2020- .................... .................... .................... .................... .................... ....................
A 8353 hCoV-19/USA/NY-WCMP9G04P/2020|EPI_ISL_427552|2020- .................... .................... .................... .................... .................... ....................
A 8354 hCoV-19/USA/NY-WCMP10A02P/2020|EPI_ISL_427553|2020 .................... .................... .................... .................... .................... ....................
A 8355 hCoV-19/USA/NY-WCMP10A05P/2020|EPI_ISL_427554|2020 .................... .................... .................... .................... .................... ....................
A 8356 hCoV-19/USA/NY-WCMP10A06P/2020|EPI_ISL_427555|2020 .................... .................... .................... .................... .................... ....................
A 8357 hCoV-19/USA/NY-WCMP10A12P/2020|EPI_ISL_427556|2020 .................... .................... .................... .................... .................... ....................
A 8358 hCoV-19/USA/NY-WCMP10B03P/2020|EPI_ISL_427557|2020 .................... .................... .................... .................... .................... ....................
A 8359 hCoV-19/USA/NY-WCMP10B07N/2020|EPI_ISL_427558|2020 .................... .................... .................... .................... .................... ....................
A 8360 hCoV-19/USA/NY-WCMP10B08P/2020|EPI_ISL_427559|2020 .................... .................... .................... .................... .................... ....................
A 8361 hCoV-19/USA/NY-WCMP10B11P/2020|EPI_ISL_427560|2020 .................... .................... .................... .................... .................... ....................
A 8362 hCoV-19/USA/NY-WCMP10B12P/2020|EPI_ISL_427561|2020 .................... .................... .................... .................... .................... ....................
A 8363 hCoV-19/USA/NY-WCMP10C03P/2020|EPI_ISL_427562|2020 .................... .................... .................... .................... .................... ....................
A 8364 hCoV-19/USA/NY-WCMP10C04P/2020|EPI_ISL_427563|2020 .................... .................... .................... .................... .................... ....................
A 8365 hCoV-19/USA/NY-WCMP10C09P/2020|EPI_ISL_427564|2020 .................... .................... .................... .................... .................... ....................
A 8366 hCoV-19/USA/NY-WCMP10C11P/2020|EPI_ISL_427565|2020 .................... .................... .................... .................... .................... ....................
A 8367 hCoV-19/USA/NY-WCMP10D06P/2020|EPI_ISL_427566|2020 .................... .................... .................... .................... .................... ....................
A 8368 hCoV-19/USA/NY-WCMP10D09P/2020|EPI_ISL_427567|2020 .................... .................... .................... .................... .................... ....................
A 8369 hCoV-19/USA/NY-WCMP10D11P/2020|EPI_ISL_427568|2020 .................... .................... .................... .................... .................... ....................

| | | | | | | | | |
|---|---|---|---|---|---|---|---|---|
| A | 8370 | hCoV-19/USA/NY-WCMP10E02P/2020\|EPI_ISL_427569\|2020 | .................... | .................... | .................... | .................... | .................... | .................... |
| A | 8371 | hCoV-19/Singapore/43/2020\|EPI_ISL_428822\|2020-02-1 | .................... | .................... | .................... | .................... | .................... | .................... |
| A | 8372 | hCoV-19/Singapore/44/2020\|EPI_ISL_428823\|2020-03-0 | .................... | .................... | .................... | .................... | .................... | .................... |
| A | 8373 | hCoV-19/Singapore/45/2020\|EPI_ISL_428824\|2020-03-1 | .................... | .................... | .................... | .................... | .................... | .................... |
| A | 8374 | hCoV-19/Singapore/46/2020\|EPI_ISL_428825\|2020-03-1 | .................... | .................... | .................... | .................... | .................... | .................... |
| A | 8375 | hCoV-19/Singapore/47/2020\|EPI_ISL_428826\|2020-03-1 | .................... | .................... | .................... | .................... | .................... | .................... |
| A | 8376 | hCoV-19/Singapore/48/2020\|EPI_ISL_428827\|2020-03-1 | .................... | .................... | .................... | .................... | .................... | .................... |
| A | 8377 | hCoV-19/Singapore/49/2020\|EPI_ISL_428828\|2020-03-0 | .................... | .................... | .................... | .................... | .................... | .................... |
| A | 8378 | hCoV-19/Singapore/50/2020\|EPI_ISL_428829\|2020-03-0 | .................... | .................... | .................... | .................... | .................... | .................... |
| A | 8379 | hCoV-19/Singapore/51/2020\|EPI_ISL_428830\|2020-02-2 | .................... | .................... | .................... | .................... | .................... | .................... |
| A | 8380 | hCoV-19/Singapore/52/2020\|EPI_ISL_428831\|2020-03-1 | .................... | .................... | .................... | .................... | .................... | .................... |
| A | 8381 | hCoV-19/Singapore/53/2020\|EPI_ISL_428832\|2020-04-0 | .................... | .................... | .................... | .................... | .................... | .................... |
| A | 8382 | hCoV-19/Singapore/54/2020\|EPI_ISL_428833\|2020-04-0 | .................... | .................... | .................... | .................... | .................... | .................... |
| A | 8383 | hCoV-19/Singapore/55/2020\|EPI_ISL_428834\|2020-04-0 | .................... | .................... | .................... | .................... | .................... | .................... |
| A | 8384 | hCoV-19/Singapore/56/2020\|EPI_ISL_428835\|2020-04-0 | .................... | .................... | .................... | .................... | .................... | .................... |
| A | 8385 | hCoV-19/Singapore/57/2020\|EPI_ISL_428836\|2020-04-0 | .................... | .................... | .................... | .................... | .................... | .................... |
| A | 8386 | hCoV-19/Singapore/58/2020\|EPI_ISL_428837\|2020-04-0 | .................... | .................... | .................... | .................... | .................... | .................... |
| A | 8387 | hCoV-19/Singapore/59/2020\|EPI_ISL_428838\|2020-04-0 | .................... | .................... | .................... | .................... | .................... | .................... |
| A | 8388 | hCoV-19/Singapore/60/2020\|EPI_ISL_428839\|2020-04-0 | .................... | .................... | .................... | .................... | .................... | .................... |
| A | 8389 | hCoV-19/Singapore/61/2020\|EPI_ISL_428840\|2020-04-0 | .................... | .................... | .................... | .................... | .................... | .................... |
| A | 8390 | hCoV-19/Singapore/62/2020\|EPI_ISL_428841\|2020-04-0 | .................... | .................... | .................... | .................... | .................... | .................... |
| A | 8391 | hCoV-19/Singapore/63/2020\|EPI_ISL_428842\|2020-04-0 | .................... | .................... | .................... | .................... | .................... | .................... |
| A | 8392 | hCoV-19/Singapore/64/2020\|EPI_ISL_428843\|2020-04-0 | .................... | .................... | .................... | .................... | .................... | .................... |
| A | 8393 | hCoV-19/Singapore/65/2020\|EPI_ISL_428844\|2020-04-0 | .................... | .................... | .................... | .................... | .................... | .................... |
| A | 8394 | hCoV-19/Singapore/66/2020\|EPI_ISL_428845\|2020-04-0 | .................... | .................... | .................... | .................... | .................... | .................... |
| A | 8395 | hCoV-19/Singapore/67/2020\|EPI_ISL_428846\|2020-04-1 | .................... | .................... | .................... | .................... | .................... | .................... |
| A | 8396 | hCoV-19/Singapore/68/2020\|EPI_ISL_428847\|2020-04-1 | .................... | .................... | .................... | .................... | .................... | .................... |
| A | 8397 | hCoV-19/Singapore/69/2020\|EPI_ISL_428848\|2020-04-1 | .................... | .................... | .................... | .................... | .................... | .................... |
| A | 8398 | hCoV-19/Singapore/70/2020\|EPI_ISL_428849\|2020-04-1 | .................... | .................... | .................... | .................... | .................... | .................... |
| A | 8399 | hCoV-19/Singapore/71/2020\|EPI_ISL_428850\|2020-04-1 | .................... | .................... | .................... | .................... | .................... | .................... |
| A | 8400 | hCoV-19/Russia/Moscow-25I/2020\|EPI_ISL_428851\|2020 | .................... | .................... | .................... | .................... | .................... | .................... |
| A | 8401 | hCoV-19/Russia/Moscow-35I/2020\|EPI_ISL_428852\|2020 | .................... | .................... | .................... | .................... | .................... | .................... |
| A | 8402 | hCoV-19/Italy/FVG-ICGEB-S18/2020\|EPI_ISL_428854\|20 | .................... | .................... | .................... | .................... | .................... | .................... |
| A | 8403 | hCoV-19/Gambia/GC19-015/2020\|EPI_ISL_428855\|2020-0 | .................... | .................... | .................... | .................... | .................... | .................... |
| A | 8404 | hCoV-19/Gambia/GC19-026/2020\|EPI_ISL_428856\|2020-0 | .................... | .................... | .................... | .................... | .................... | .................... |
| A | 8405 | hCoV-19/Gambia/GC19-029/2020\|EPI_ISL_428857\|2020-0 | .................... | .................... | .................... | .................... | .................... | .................... |
| A | 8406 | hCoV-19/Russia/Moscow-62505/2020\|EPI_ISL_428860\|20 | .................... | .................... | .................... | .................... | .................... | .................... |
| A | 8407 | hCoV-19/Russia/Moscow-62501/2020\|EPI_ISL_428861\|20 | .................... | .................... | .................... | .................... | .................... | .................... |
| A | 8408 | hCoV-19/Russia/Moscow-62506/2020\|EPI_ISL_428862\|20 | .................... | .................... | .................... | .................... | .................... | .................... |
| A | 8409 | hCoV-19/Russia/Moscow-62508/2020\|EPI_ISL_428863\|20 | .................... | .................... | .................... | .................... | .................... | .................... |
| A | 8410 | hCoV-19/Russia/Moscow-62502/2020\|EPI_ISL_428864\|20 | .................... | .................... | .................... | .................... | .................... | .................... |
| A | 8411 | hCoV-19/Russia/Lipetsk-62704/2020\|EPI_ISL_428865\|2 | .................... | .................... | .................... | .................... | .................... | .................... |
| A | 8412 | hCoV-19/Russia/Lipetsk-62707/2020\|EPI_ISL_428866\|2 | .................... | .................... | .................... | .................... | .................... | .................... |
| A | 8413 | hCoV-19/Russia/Krasnodar-63401/2020\|EPI_ISL_428867 | .................... | .................... | .................... | .................... | .................... | .................... |
| A | 8414 | hCoV-19/Russia/StPetersburg-64304/2020\|EPI_ISL_428 | .................... | .................... | .................... | .................... | .................... | .................... |
| A | 8415 | hCoV-19/Russia/Moscow_64405/2020\|EPI_ISL_428869\|20 | .................... | .................... | .................... | .................... | .................... | .................... |
| A | 8416 | hCoV-19/Russia/Moscow-64601/2020\|EPI_ISL_428870\|20 | .................... | .................... | .................... | .................... | .................... | .................... |
| A | 8417 | hCoV-19/Russia/Krasnoyarsk-66501/2020\|EPI_ISL_4288 | .................... | .................... | .................... | .................... | .................... | .................... |
| A | 8418 | hCoV-19/Russia/Moscow-71605/2020\|EPI_ISL_428872\|20 | .................... | .................... | .................... | .................... | .................... | .................... |
| A | 8419 | hCoV-19/Russia/Sverdlovsk-71904/2020\|EPI_ISL_42887 | .................... | .................... | .................... | .................... | .................... | .................... |
| A | 8420 | hCoV-19/Russia/Yakutia-73709/2020\|EPI_ISL_428874\|2 | .................... | .................... | .................... | .................... | .................... | .................... |
| A | 8421 | hCoV-19/Russia/Moscow-67117/2020\|EPI_ISL_428875\|20 | .................... | .................... | .................... | .................... | .................... | .................... |
| A | 8422 | hCoV-19/Russia/Moscow-71604/2020\|EPI_ISL_428876\|20 | .................... | .................... | .................... | .................... | .................... | .................... |
| A | 8423 | hCoV-19/Russia/Moscow-71602/2020\|EPI_ISL_428877\|20 | .................... | .................... | .................... | .................... | .................... | .................... |
| A | 8424 | hCoV-19/Russia/Omsk-89001/2020\|EPI_ISL_428878\|2020 | .................... | .................... | .................... | .................... | .................... | .................... |
| A | 8425 | hCoV-19/Russia/Sverdlovsk-71905/2020\|EPI_ISL_42887 | .................... | .................... | .................... | .................... | .................... | .................... |
| A | 8426 | hCoV-19/Russia/Khabarovsk-71403/2020\|EPI_ISL_42888 | .................... | .................... | .................... | .................... | .................... | .................... |
| A | 8427 | hCoV-19/Russia/Moscow-67142/2020\|EPI_ISL_428881\|20 | .................... | .................... | .................... | .................... | .................... | .................... |
| A | 8428 | hCoV-19/Russia/Moscow-67609/2020\|EPI_ISL_428882\|20 | .................... | .................... | .................... | .................... | .................... | .................... |
| A | 8429 | hCoV-19/Russia/StPetersburg-73603/2020\|EPI_ISL_428 | .................... | .................... | .................... | .................... | .................... | .................... |
| A | 8430 | hCoV-19/Russia/Moscow-77627/2020\|EPI_ISL_428884\|20 | .................... | .................... | .................... | .................... | .................... | .................... |
| A | 8431 | hCoV-19/Russia/Samara-73406/2020\|EPI_ISL_428885\|20 | .................... | .................... | .................... | .................... | .................... | .................... |
| A | 8432 | hCoV-19/Russia/Samara-73403/2020\|EPI_ISL_428886\|20 | .................... | .................... | .................... | .................... | .................... | .................... |
| A | 8433 | hCoV-19/Russia/Moscow-77610/2020\|EPI_ISL_428887\|20 | .................... | .................... | .................... | .................... | .................... | .................... |
| A | 8434 | hCoV-19/Russia/Yakutia-73703/2020\|EPI_ISL_428888\|2 | .................... | .................... | .................... | .................... | .................... | .................... |
| A | 8435 | hCoV-19/Russia/Moscow-77620/2020\|EPI_ISL_428889\|20 | .................... | .................... | .................... | .................... | .................... | .................... |
| A | 8436 | hCoV-19/Russia/Sverdlovsk-71903/2020\|EPI_ISL_42889 | .................... | .................... | .................... | .................... | .................... | .................... |
| A | 8437 | hCoV-19/Russia/Yakutia-83301/2020\|EPI_ISL_428891\|2 | .................... | .................... | .................... | .................... | .................... | .................... |
| A | 8438 | hCoV-19/Russia/Krasnodar-80902/2020\|EPI_ISL_428892 | .................... | .................... | .................... | .................... | .................... | .................... |
| A | 8439 | hCoV-19/Russia/Kabardino-Balkaria-80501/2020\|EPI_I | .................... | .................... | .................... | .................... | .................... | .................... |
| A | 8440 | hCoV-19/Russia/Buryatia-84506/2020\|EPI_ISL_428894\| | .................... | .................... | .................... | .................... | .................... | .................... |
| A | 8441 | hCoV-19/Russia/Perm-82707/2020\|EPI_ISL_428895\|2020 | .................... | .................... | .................... | .................... | .................... | .................... |
| A | 8442 | hCoV-19/Russia/Vector_84801/2020\|EPI_ISL_428896\|20 | .................... | .................... | .................... | .................... | .................... | .................... |
| A | 8443 | hCoV-19/Russia/StPetersburg-82005/2020\|EPI_ISL_428 | .................... | ............A....... | .................... | .................... | .................... | .................... |
| A | 8444 | hCoV-19/Russia/Kabardino-Balkaria-80503/2020\|EPI_I | .................... | .................... | .................... | .................... | .................... | .................... |
| A | 8445 | hCoV-19/Russia/Buryatia-86204/2020\|EPI_ISL_428899\| | .................... | .................... | .................... | .................... | .................... | .................... |
| A | 8446 | hCoV-19/USA/KY-9A06/2020\|EPI_ISL_468063\|2020-03-18 | .................... | .................... | .................... | .................... | .................... | .................... |
| A | 8447 | hCoV-19/USA/KY-9A10/2020\|EPI_ISL_468064\|2020-03-18 | .................... | .................... | .................... | .................... | .................... | .................... |

| | | | | | | | | |
|---|---|---|---|---|---|---|---|---|
| A | 8448 hCoV-19/USA/KY-9E09/2020\|EPI_ISL_468065\|2020-03-18 | ···················· | ···················· | ····················· | ···················· | ···················· | ···················· | |
| A | 8449 hCoV-19/England/01/2020\|EPI_ISL_407071\|2020-01-29 | ···················· | ···················· | ····················· | ···················· | ···················· | ···················· | |
| A | 8450 hCoV-19/England/02/2020\|EPI_ISL_407073\|2020-01-29 | ···················· | ···················· | ····················· | ···················· | ···················· | ···················· | |
| A | 8451 hCoV-19/Finland/1/2020\|EPI_ISL_407079\|2020-01-29 | ···················· | ···················· | ····················· | ···················· | ···················· | ···················· | |
| A | 8452 hCoV-19/USA/WA1-A12/2020\|EPI_ISL_407214\|2020-01-25 | ···················· | ···················· | ····················· | ···················· | ···················· | ···················· | |
| A | 8453 hCoV-19/USA/WA1-F6/2020\|EPI_ISL_407215\|2020-01-25 | ···················· | ···················· | ····················· | ···················· | ···················· | ···················· | |
| A | 8454 hCoV-19/Netherlands/NoordBrabant_42/2020\|EPI_ISL_4 | ···················· | ···················· | ····················· | ···················· | ···················· | ···················· | |
| A | 8455 hCoV-19/Netherlands/NoordBrabant_44/2020\|EPI_ISL_4 | ···················· | ···················· | ····················· | ···················· | ···················· | ···················· | |
| A | 8456 hCoV-19/Netherlands/NoordBrabant_45/2020\|EPI_ISL_4 | ···················· | ···················· | ····················· | ···················· | ···················· | ···················· | |
| A | 8457 hCoV-19/Netherlands/NoordBrabant_46/2020\|EPI_ISL_4 | ···················· | ···················· | ····················· | ···················· | ···················· | ···················· | |
| A | 8458 hCoV-19/Netherlands/NoordBrabant_47/2020\|EPI_ISL_4 | ···················· | ···················· | ····················· | ···················· | ···················· | ···················· | |
| A | 8459 hCoV-19/Malaysia/MKAK-CL-2020-5045/2020\|EPI_ISL_41 | ···················· | ···················· | ····················· | ···················· | ···················· | ···················· | |
| A | 8460 hCoV-19/USA/NY-NYUMC2/2020\|EPI_ISL_416830\|2020-03- | ···················· | ···················· | ····················· | ···················· | ···················· | ···················· | |
| A | 8461 hCoV-19/USA/NY-NYUMC3/2020\|EPI_ISL_416831\|2020-03- | ···················· | ···················· | ····················· | ···················· | ···················· | ···················· | |
| A | 8462 hCoV-19/USA/NY-NYUMC4/2020\|EPI_ISL_416832\|2020-03- | ···················· | ···················· | ····················· | ···················· | ···················· | ···················· | |
| A | 8463 hCoV-19/Israel/ISR_JP0320/2020\|EPI_ISL_419211\|2020 | ···················· | ···················· | ····················· | ···················· | ···················· | ···················· | |
| A | 8464 hCoV-19/Hong Kong/HKPU39-3001/2020\|EPI_ISL_419214\| | ···················· | ···················· | ····················· | ···················· | ···················· | ···················· | |
| A | 8465 hCoV-19/Hong Kong/HKPU40-2801/2020\|EPI_ISL_419215\| | ···················· | ···················· | ····················· | ···················· | ···················· | ···················· | |
| A | 8466 hCoV-19/Hong Kong/HKPU41-0802/2020\|EPI_ISL_419216\| | ···················· | ···················· | ····················· | ···················· | ···················· | ···················· | |
| A | 8467 hCoV-19/Hong Kong/HKPU42-0302/2020\|EPI_ISL_419217\| | ···················· | ···················· | ····················· | ···················· | ···················· | ···················· | |
| A | 8468 hCoV-19/Hong Kong/HKPU44-0102/2020\|EPI_ISL_419219\| | ···················· | ···················· | ····················· | ···················· | ···················· | ···················· | |
| A | 8469 hCoV-19/Hong Kong/HKPU45-0502/2020\|EPI_ISL_419221\| | ···················· | ···················· | ····················· | ···················· | ···················· | ···················· | |
| A | 8470 hCoV-19/Hong Kong/HKPU48-0202/2020\|EPI_ISL_419222\| | ···················· | ···················· | ····················· | ···················· | ···················· | ···················· | |
| A | 8471 hCoV-19/Hong Kong/HKPU52-3101/2020\|EPI_ISL_419224\| | ···················· | ···················· | ····················· | ···················· | ···················· | ···················· | |
| A | 8472 hCoV-19/Hong Kong/HKPU53-0802/2020\|EPI_ISL_419225\| | ···················· | ···················· | ····················· | ···················· | ···················· | ···················· | |
| A | 8473 hCoV-19/Hong Kong/HKPU54-0302/2020\|EPI_ISL_419226\| | ···················· | ···················· | ····················· | ···················· | ···················· | ··················C· | 1 |
| A | 8474 hCoV-19/Hong Kong/HKPU57-0702/2020\|EPI_ISL_419227\| | ···················· | ···················· | ····················· | ···················· | ···················· | ···················· | |
| A | 8475 hCoV-19/Hong Kong/HKPU60-0802/2020\|EPI_ISL_419228\| | ···················· | ···················· | ····················· | ···················· | ···················· | ···················· | |
| A | 8476 hCoV-19/Hong Kong/HKPU61-0202/2020\|EPI_ISL_419229\| | ···················· | ···················· | ····················· | ···················· | ···················· | ···················· | |
| A | 8477 hCoV-19/Spain/AN-ISCIII-201617/2020\|EPI_ISL_419230 | ···················· | ···················· | ····················· | ···················· | ···················· | ···················· | |
| A | 8478 hCoV-19/Hong Kong/HKPU63-1402/2020\|EPI_ISL_419231\| | ···················· | ···················· | ····················· | ···················· | ···················· | ···················· | |
| A | 8479 hCoV-19/Hong Kong/HKPU64-1202/2020\|EPI_ISL_419232\| | ···················· | ···················· | ····················· | ···················· | ···················· | ···················· | |
| A | 8480 hCoV-19/Spain/CN-ISCIII-201495/2020\|EPI_ISL_419233 | ···················· | ···················· | ····················· | ···················· | ···················· | ···················· | |
| A | 8481 hCoV-19/Spain/RI-ISCIII-201575/2020\|EPI_ISL_419234 | ···················· | ···················· | ····················· | ···················· | ···················· | ···················· | |
| A | 8482 hCoV-19/Spain/MD-ISCIII-201706/2020\|EPI_ISL_419235 | ···················· | ···················· | ····················· | ···················· | ···················· | ···················· | |
| A | 8483 hCoV-19/Spain/MD-ISCIII-201709/2020\|EPI_ISL_419236 | ···················· | ···················· | ····················· | ···················· | ···················· | ···················· | |
| A | 8484 hCoV-19/Spain/MD-ISCIII-201738/2020\|EPI_ISL_419237 | ···················· | ···················· | ····················· | ···················· | ···················· | ···················· | |
| A | 8485 hCoV-19/Spain/PV-ISCIII-201493/2020\|EPI_ISL_419238 | ···················· | ···················· | ····················· | ···················· | ···················· | ···················· | |
| A | 8486 hCoV-19/Spain/PV-ISCIII-201607/2020\|EPI_ISL_419240 | ···················· | ···················· | ····················· | ···················· | ···················· | ···················· | |
| A | 8487 hCoV-19/Hong Kong/HKPU74-1302/2020\|EPI_ISL_419245\| | ···················· | ···················· | ····················· | ···················· | ···················· | ···················· | |
| A | 8488 hCoV-19/Hong Kong/HKPU83-1702/2020\|EPI_ISL_419247\| | ···················· | ···················· | ····················· | ···················· | ···················· | ···················· | |
| A | 8489 hCoV-19/USA/VA-DCLS-00010/2020\|EPI_ISL_419262\|2020 | ···················· | ···················· | ····················· | ···················· | ···················· | ···················· | |
| A | 8490 hCoV-19/USA/VA-DCLS-00011/2020\|EPI_ISL_419263\|2020 | ···················· | ···················· | ····················· | ···················· | ···················· | ···················· | |
| A | 8491 hCoV-19/Belgium/UGent-12/2020\|EPI_ISL_419264\|2020- | ···················· | ···················· | ····················· | ···················· | ···················· | ···················· | |
| A | 8492 hCoV-19/Belgium/UGent-6/2020\|EPI_ISL_419265\|2020-0 | ···················· | ···················· | ····················· | ···················· | ···················· | ···················· | |
| A | 8493 hCoV-19/Australia/VIC324/2020\|EPI_ISL_420015\|2020- | ···················· | ···················· | ····················· | ···················· | ···················· | ···················· | |
| A | 8494 hCoV-19/Australia/VIC325/2020\|EPI_ISL_420016\|2020- | ···················· | ···················· | ····················· | ···················· | ···················· | ···················· | |
| A | 8495 hCoV-19/Australia/VIC326/2020\|EPI_ISL_420017\|2020- | ···················· | ···················· | ····················· | ···················· | ···················· | ···················· | |
| A | 8496 hCoV-19/USA/VA-DCLS-0031/2020\|EPI_ISL_420018\|2020- | ···················· | ···················· | ····················· | ···················· | ···················· | ···················· | |
| A | 8497 hCoV-19/Algeria/G0860_2262/2020\|EPI_ISL_420037\|202 | ···················· | ···················· | ····················· | ···················· | ···················· | ···················· | |
| A | 8498 hCoV-19/France/BFC-2709/2020\|EPI_ISL_420038\|2020-0 | ···················· | ···················· | ····················· | ···················· | ···················· | ···················· | |
| A | 8499 hCoV-19/France/GES-2720/2020\|EPI_ISL_420039\|2020-0 | ···················· | ···················· | ····················· | ···················· | ···················· | ···················· | |
| A | 8500 hCoV-19/France/GES-2722/2020\|EPI_ISL_420040\|2020-0 | ···················· | ···················· | ····················· | ···················· | ···················· | ···················· | |
| A | 8501 hCoV-19/France/HDF-2748/2020\|EPI_ISL_420041\|2020-0 | ···················· | ···················· | ····················· | ···················· | ···················· | ···················· | |
| A | 8502 hCoV-19/France/IDF-2768/2020\|EPI_ISL_420042\|2020-0 | ···················· | ···················· | ····················· | ···················· | ···················· | ···················· | |
| A | 8503 hCoV-19/France/IDF-2792/2020\|EPI_ISL_420043\|2020-0 | ···················· | ···················· | ····················· | ···················· | ···················· | ···················· | |
| A | 8504 hCoV-19/France/HDF-2797/2020\|EPI_ISL_420044\|2020-0 | ···················· | ···················· | ····················· | ···················· | ···················· | ···················· | |
| A | 8505 hCoV-19/France/GES-2843/2020\|EPI_ISL_420045\|2020-0 | ···················· | ···················· | ····················· | ···················· | ···················· | ···················· | |
| A | 8506 hCoV-19/France/IDF-2848/2020\|EPI_ISL_420046\|2020-0 | ···················· | ···················· | ····················· | ···················· | ···················· | ···················· | |
| A | 8507 hCoV-19/France/IDF-2849/2020\|EPI_ISL_420047\|2020-0 | ···················· | ···················· | ····················· | ···················· | ···················· | ···················· | |
| A | 8508 hCoV-19/France/IDF-2936/2020\|EPI_ISL_420048\|2020-0 | ···················· | ···················· | ····················· | ···················· | ···················· | ···················· | |
| A | 8509 hCoV-19/France/HDF-2946/2020\|EPI_ISL_420049\|2020-0 | ···················· | ···················· | ····················· | ···················· | ···················· | ···················· | |
| A | 8510 hCoV-19/France/HDF-2948/2020\|EPI_ISL_420050\|2020-0 | ···················· | ···················· | ····················· | ···················· | ···················· | ···················· | |
| A | 8511 hCoV-19/France/IDF-2989/2020\|EPI_ISL_420051\|2020-0 | ···················· | ···················· | ····················· | ···················· | ···················· | ···················· | |
| A | 8512 hCoV-19/France/IDF-3029/2020\|EPI_ISL_420052\|2020-0 | ···················· | ···················· | ····················· | ···················· | ···················· | ···················· | |
| A | 8513 hCoV-19/France/HDF-3036/2020\|EPI_ISL_420053\|2020-0 | ···················· | ···················· | ····················· | ···················· | ···················· | ···················· | |
| A | 8514 hCoV-19/France/GES-3067/2020\|EPI_ISL_420055\|2020-0 | ···················· | ···················· | ····················· | ···················· | ···················· | ···················· | |
| A | 8515 hCoV-19/France/HDF-3138/2020\|EPI_ISL_420056\|2020-0 | ···················· | ···················· | ····················· | ···················· | ···················· | ···················· | |
| A | 8516 hCoV-19/France/HDF-3141/2020\|EPI_ISL_420057\|2020-0 | ···················· | ···················· | ··A·················· | ···················· | ···················· | ···················· | |
| A | 8517 hCoV-19/France/IDF-3163/2020\|EPI_ISL_420058\|2020-0 | ···················· | ···················· | ····················· | ···················· | ···················· | ···················· | |
| A | 8518 hCoV-19/France/IDF-3165/2020\|EPI_ISL_420059\|2020-0 | ···················· | ···················· | ····················· | ···················· | ···················· | ···················· | |
| A | 8519 hCoV-19/France/IDF-3170/2020\|EPI_ISL_420060\|2020-0 | ···················· | ···················· | ····················· | ···················· | ···················· | ···················· | |
| A | 8520 hCoV-19/France/IDF-3212/2020\|EPI_ISL_420061\|2020-0 | ···················· | ···················· | ····················· | ···················· | ···················· | ···················· | |
| A | 8521 hCoV-19/France/IDF-3230/2020\|EPI_ISL_420063\|2020-0 | ···················· | ···················· | ····················· | ···················· | ···················· | ···················· | |
| A | 8522 hCoV-19/France/IDF-3235/2020\|EPI_ISL_420064\|2020-0 | ···················· | ···················· | ····················· | ···················· | ···················· | ···················· | |
| A | 8523 hCoV-19/Estonia/ChVir1982/2020\|EPI_ISL_420065\|2020 | ···················· | ···················· | ····················· | ···················· | ···················· | ···················· | |
| A | 8524 hCoV-19/Estonia/ChVir1983/2020\|EPI_ISL_420066\|2020 | ···················· | ···················· | ····················· | ···················· | ···················· | ···················· | |
| A | 8525 hCoV-19/Estonia/ChVir1985/2020\|EPI_ISL_420067\|2020 | ···················· | ···················· | ····················· | ···················· | ···················· | ···················· | |

| | | | | | | | | |
|---|---|---|---|---|---|---|---|---|
| A | 8526 hCoV-19/Senegal/306/2020\|EPI_ISL_420069\|2020-03-17 | ···················· | ···················· | ···················· | ···················· | ···················· | ···················· | |
| A | 8527 hCoV-19/Senegal/315/2020\|EPI_ISL_420070\|2020-03-17 | ···················· | ···················· | ···················· | ···················· | ···················· | ···················· | |
| A | 8528 hCoV-19/Senegal/370/2020\|EPI_ISL_420072\|2020-03-18 | ···················· | ···················· | ···················· | ···················· | ···················· | ···················· | |
| A | 8529 hCoV-19/Senegal/382/2020\|EPI_ISL_420073\|2020-03-19 | ···················· | ···················· | ···················· | ···················· | ···················· | ···················· | |
| A | 8530 hCoV-19/Senegal/600/2020\|EPI_ISL_420074\|2020-03-20 | ···················· | ···················· | ···················· | ···················· | ···················· | ···················· | |
| A | 8531 hCoV-19/Senegal/611/2020\|EPI_ISL_420076\|2020-03-20 | ···················· | ···················· | ···················· | ···················· | ···················· | ···················· | |
| A | 8532 hCoV-19/Senegal/618/2020\|EPI_ISL_420077\|2020-03-20 | ···················· | ···················· | ···················· | ···················· | ···················· | ···················· | |
| A | 8533 hCoV-19/Senegal/620/2020\|EPI_ISL_420078\|2020-03-20 | ···················· | ···················· | ···················· | ···················· | ···················· | ···················· | |
| A | 8534 hCoV-19/Russia/StPetersburg-RII3992/2020\|EPI_ISL_4 | ···················· | ···················· | ···················· | ···················· | ···················· | ···················· | |
| A | 8535 hCoV-19/Russia/StPetersburg-RII3997/2020\|EPI_ISL_4 | ···················· | ···················· | ···················· | ···················· | ···················· | ···················· | |
| A | 8536 hCoV-19/Taiwan/125/2020\|EPI_ISL_420082\|2020-03-19 | ···················· | ···················· | ···················· | ···················· | ···················· | ···················· | |
| A | 8537 hCoV-19/Taiwan/128/2020\|EPI_ISL_420083\|2020-03-18 | ···················· | ···················· | ···················· | ···················· | ···················· | ···················· | |
| A | 8538 hCoV-19/Taiwan/170/2020\|EPI_ISL_420084\|2020-03-21 | ···················· | ···················· | ···················· | ···················· | ···················· | ···················· | |
| A | 8539 hCoV-19/Taiwan/107/2020\|EPI_ISL_420085\|2020-03-20 | ···················· | ···················· | ···················· | ···················· | ···················· | ···················· | |
| A | 8540 hCoV-19/Singapore/22/2020\|EPI_ISL_420099\|2020-03-0 | ···················· | ···················· | ···················· | ···················· | ···················· | ···················· | |
| A | 8541 hCoV-19/England/SHEF-C04C4/2020\|EPI_ISL_420200\|202 | ···················· | ···················· | ···················· | ···················· | ···················· | ···················· | |
| A | 8542 hCoV-19/England/SHEF-C0488/2020\|EPI_ISL_420201\|202 | ···················· | ···················· | ···················· | ···················· | ···················· | ···················· | |
| A | 8543 hCoV-19/England/SHEF-C04B5/2020\|EPI_ISL_420202\|202 | ···················· | ···················· | ···················· | ···················· | ···················· | ···················· | |
| A | 8544 hCoV-19/England/SHEF-C045B/2020\|EPI_ISL_420203\|202 | ···················· | ···················· | ···················· | ···················· | ···················· | ···················· | |
| A | 8545 hCoV-19/England/SHEF-C0215/2020\|EPI_ISL_420204\|202 | ···················· | ···················· | ···················· | ···················· | ···················· | ···················· | |
| A | 8546 hCoV-19/England/SHEF-C034F/2020\|EPI_ISL_420205\|202 | ···················· | ···················A | ···················· | ···················· | ···················· | ···················· | 1 |
| A | 8549 hCoV-19/England/SHEF-C02AC/2020\|EPI_ISL_420208\|202 | ···················· | ······A············· | ···················· | ···················· | ···················· | ···················· | |
| A | 8550 hCoV-19/England/SHEF-C027F/2020\|EPI_ISL_420209\|202 | ···················· | ···················· | ···················· | ···················· | ···················· | ···················· | |
| A | 8551 hCoV-19/England/SHEF-C041F/2020\|EPI_ISL_420210\|202 | ···················· | ···················· | ···················· | ···················· | ···················· | ···················· | |
| A | 8552 hCoV-19/England/SHEF-C0224/2020\|EPI_ISL_420211\|202 | ···················· | ···················· | ···················· | ···················· | ···················· | ···················· | |
| A | 8553 hCoV-19/England/SHEF-C03D6/2020\|EPI_ISL_420212\|202 | ···················· | ···················· | ···················· | ···················· | ···················· | ···················· | |
| A | 8554 hCoV-19/England/SHEF-C04D3/2020\|EPI_ISL_420213\|202 | ···················· | ···················· | ···················· | ···················· | ···················· | ···················· | |
| A | 8556 hCoV-19/England/SHEF-C046A/2020\|EPI_ISL_420215\|202 | ···················· | ···················· | ···················· | ···················· | ···················· | ···················· | |
| A | 8557 hCoV-19/England/SHEF-C0312/2020\|EPI_ISL_420216\|202 | ···················· | ···················· | ···················· | ···················· | ···················· | ···················· | |
| A | 8558 hCoV-19/England/SHEF-C043D/2020\|EPI_ISL_420217\|202 | ···················· | ···················· | ···················· | ···················· | ···················· | ···················· | |
| A | 8560 hCoV-19/England/SHEF-C037C/2020\|EPI_ISL_420219\|202 | ···················· | ···················· | ···················· | ···················· | ···················· | ···················· | |
| A | 8561 hCoV-19/England/SHEF-C035E/2020\|EPI_ISL_420220\|202 | ···················· | ···················· | ···················· | ···················· | ···················· | ···················· | |
| A | 8562 hCoV-19/England/SHEF-C03E5/2020\|EPI_ISL_420221\|202 | ···················· | ···················· | ···················· | ···················· | ···················· | ···················· | |
| A | 8563 hCoV-19/England/SHEF-C02F7/2020\|EPI_ISL_420222\|202 | ···················· | ···················· | ···················· | ···················· | ···················· | ···················· | |
| A | 8564 hCoV-19/England/SHEF-C0716/2020\|EPI_ISL_420223\|202 | ···················· | ···················· | ···················· | ···················· | ···················· | ···················· | |
| A | 8565 hCoV-19/England/SHEF-C0594/2020\|EPI_ISL_420224\|202 | ···················· | ···················· | ···················· | ···················· | ···················· | ···················· | |
| A | 8566 hCoV-19/England/SHEF-C052B/2020\|EPI_ISL_420225\|202 | ···················· | ···················· | ···················· | ···················· | ···················· | ···················· | |
| A | 8567 hCoV-19/England/SHEF-C0154/2020\|EPI_ISL_420226\|202 | ···················· | ···················· | ···················· | ···················· | ···················· | ···················· | |
| A | 8568 hCoV-19/England/SHEF-C01FA/2020\|EPI_ISL_420227\|202 | ···················· | ···················· | ···················· | ···················· | ···················· | ···················· | |
| A | 8569 hCoV-19/England/SHEF-C0127/2020\|EPI_ISL_420228\|202 | ···················· | ···················· | ···················· | ···················· | ···················· | ···················· | |
| A | 8570 hCoV-19/England/SHEF-C01CD/2020\|EPI_ISL_420229\|202 | ···················· | ···················· | ···················· | ···················· | ···················· | ···················· | |
| A | 8571 hCoV-19/England/SHEF-C0145/2020\|EPI_ISL_420230\|202 | ···················· | ···················· | ···················· | ···················· | ···················· | ···················· | |
| A | 8572 hCoV-19/England/SHEF-C0691/2020\|EPI_ISL_420231\|202 | ···················· | ···················· | ···················· | ···················· | ···················· | ···················· | |
| A | 8573 hCoV-19/England/SHEF-C0682/2020\|EPI_ISL_420232\|202 | ···················· | ···················· | ···················· | ···················· | ···················· | ···················· | |
| A | 8574 hCoV-19/England/SHEF-C050D/2020\|EPI_ISL_420233\|202 | ···················· | ···················· | ···················· | ···················· | ···················· | ···················· | |
| A | 8575 hCoV-19/England/SHEF-C0567/2020\|EPI_ISL_420234\|202 | ···················· | ···················· | ···················· | ···················· | ···················· | ···················· | |
| A | 8576 hCoV-19/England/SHEF-C01BE/2020\|EPI_ISL_420253\|202 | ···················· | ···················· | ···················· | ···················· | ···················· | ···················· | |
| A | 8577 hCoV-19/England/SHEF-C01EB/2020\|EPI_ISL_420254\|202 | ···················· | ···················· | ···················· | ···················· | ···················· | ···················· | |
| A | 8578 hCoV-19/England/SHEF-C0163/2020\|EPI_ISL_420255\|202 | ···················· | ···················· | ···················· | ···················· | ···················· | ···················· | |
| A | 8579 hCoV-19/England/SHEF-C01DC/2020\|EPI_ISL_420256\|202 | ···················· | ···················· | ···················· | ···················· | ···················· | ···················· | |
| A | 8580 hCoV-19/USA/WY-WYPHL004/2020\|EPI_ISL_421346\|2020-0 | ···················· | ···················· | ···················· | ···················· | ···················· | ···················· | |
| A | 8581 hCoV-19/USA/NY-PV08109/2020\|EPI_ISL_421348\|2020-03 | ···················· | ···················· | ···················· | ···················· | ···················· | ···················· | |
| A | 8582 hCoV-19/USA/NY-PV08120/2020\|EPI_ISL_421349\|2020-03 | ···················· | ···················· | ···················· | ···················· | ···················· | ···················· | |
| A | 8583 hCoV-19/USA/NY-PV08401/2020\|EPI_ISL_421369\|2020-03 | ···················· | ···················· | ···················· | ···················· | ···················· | ···················· | |
| A | 8584 hCoV-19/USA/NY-PV08402/2020\|EPI_ISL_421370\|2020-03 | ···················· | ···················· | ···················· | ···················· | ···················· | ···················· | |
| A | 8585 hCoV-19/USA/NY-PV08403/2020\|EPI_ISL_421371\|2020-03 | ···················· | ···················· | ···················· | ···················· | ···················· | ···················· | |
| A | 8586 hCoV-19/USA/NY-PV08412/2020\|EPI_ISL_421375\|2020-03 | ···················· | ···················· | ···················· | ···················· | ···················· | ···················· | |
| A | 8587 hCoV-19/USA/NY-PV08413/2020\|EPI_ISL_421376\|2020-03 | ···················· | ···················· | ···················· | ···················· | ···················· | ···················· | |
| A | 8588 hCoV-19/USA/NY-PV08415/2020\|EPI_ISL_421378\|2020-03 | ···················· | ···················· | ···················· | ···················· | ···················· | ···················· | |
| A | 8589 hCoV-19/USA/NY-PV08416/2020\|EPI_ISL_421379\|2020-03 | ···················· | ···················· | ···················· | ···················· | ···················· | ···················· | |
| A | 8590 hCoV-19/USA/NY-PV08425/2020\|EPI_ISL_421383\|2020-03 | ···················· | ···················· | ···················· | ···················· | ···················· | ···················· | |
| A | 8591 hCoV-19/USA/NY-PV08426/2020\|EPI_ISL_421384\|2020-03 | ···················· | ···················· | ···················· | ···················· | ···················· | ···················· | |
| A | 8592 hCoV-19/USA/NY-PV08427/2020\|EPI_ISL_421385\|2020-03 | ···················· | ···················· | ···················· | ···················· | ···················· | ···················· | |
| A | 8593 hCoV-19/USA/NY-PV08428/2020\|EPI_ISL_421386\|2020-03 | ···················· | ···················· | ···················· | ···················· | ···················· | ···················· | |
| A | 8594 hCoV-19/USA/NY-PV08429/2020\|EPI_ISL_421387\|2020-03 | ···················· | ···················· | ···················· | ···················· | ···················· | ···················· | |
| A | 8595 hCoV-19/USA/NY-PV08432/2020\|EPI_ISL_421388\|2020-03 | ···················· | ···················· | ···················· | ···················· | ···················· | ···················· | |
| A | 8596 hCoV-19/USA/NY-PV08434/2020\|EPI_ISL_421389\|2020-03 | ···················· | ···················· | ···················· | ···················· | ···················· | ···················· | |
| A | 8597 hCoV-19/USA/NY-PV08435/2020\|EPI_ISL_421390\|2020-03 | ···················· | ···················· | ···················· | ···················· | ···················· | ···················· | |
| A | 8598 hCoV-19/USA/NY-PV08436/2020\|EPI_ISL_421391\|2020-03 | ···················· | ···················· | ···················· | ···················· | ···················· | ···················· | |
| A | 8599 hCoV-19/USA/NY-PV08443/2020\|EPI_ISL_421394\|2020-03 | ···················· | ······A············· | ···················· | ···················· | ···················· | ···················· | |
| A | 8600 hCoV-19/USA/NY-PV08444/2020\|EPI_ISL_421395\|2020-03 | ···················· | ···················· | ···················· | ···················· | ···················· | ···················· | |
| A | 8601 hCoV-19/USA/NY-PV08445/2020\|EPI_ISL_421396\|2020-03 | ···················· | ···················· | ···················· | ···················· | ···················· | ···················· | |
| A | 8602 hCoV-19/USA/NY-PV08446/2020\|EPI_ISL_421397\|2020-03 | ···················· | ···················· | ···················· | ···················· | ···················· | ···················· | |
| A | 8603 hCoV-19/USA/NY-PV08447/2020\|EPI_ISL_421398\|2020-03 | ···················· | ···················· | ···················· | ···················· | ···················· | ···················· | |
| A | 8604 hCoV-19/USA/NY-PV08448/2020\|EPI_ISL_421399\|2020-03 | ···················· | ···················· | ···················· | ···················· | ···················· | ···················· | |
| A | 8605 hCoV-19/France/HDF-2306/2020\|EPI_ISL_421500\|2020-0 | ···················· | ···················· | ···················· | ···················· | ···················· | ···················· | |
| A | 8606 hCoV-19/France/IDF-2359/2020\|EPI_ISL_421501\|2020-0 | ···················· | ···················· | ···················· | ···················· | ···················· | ···················· | |
| A | 8607 hCoV-19/France/IDF-2412/2020\|EPI_ISL_421502\|2020-0 | ···················· | ···················· | ···················· | ···················· | ···················· | ···················· | |

| | | | | | | | |
|---|---|---|---|---|---|---|---|
| A | 8608 hCoV-19/France/IDF-2414/2020\|EPI_ISL_421503\|2020-0 | ···················· | ···················· | ···················· | ···················· | ···················· | ···················· |
| A | 8609 hCoV-19/France/IDF-2536/2020\|EPI_ISL_421504\|2020-0 | ···················· | ···················· | ···················· | ···················· | ···················· | ···················· |
| A | 8610 hCoV-19/France/IDF-2548/2020\|EPI_ISL_421505\|2020-0 | ···················· | ···················· | ···················· | ···················· | ···················· | ···················· |
| A | 8611 hCoV-19/France/IDF-3236/2020\|EPI_ISL_421506\|2020-0 | ···················· | ···················· | ···················· | ···················· | ···················· | ···················· |
| A | 8612 hCoV-19/France/IDF-3274/2020\|EPI_ISL_421507\|2020-0 | ···················· | ···················· | ···················· | ···················· | ···················· | ···················· |
| A | 8613 hCoV-19/France/IDF-3276/2020\|EPI_ISL_421508\|2020-0 | ···················· | ···················· | ···················· | ···················· | ···················· | ···················· |
| A | 8614 hCoV-19/France/HDF-3290/2020\|EPI_ISL_421509\|2020-0 | ···················· | ···················· | ···················· | ···················· | ···················· | ···················· |
| A | 8615 hCoV-19/France/HDF-3293/2020\|EPI_ISL_421510\|2020-0 | ···················· | ···················· | ···················· | ···················· | ···················· | ···················· |
| A | 8616 hCoV-19/France/HDF-3295/2020\|EPI_ISL_421511\|2020-0 | ···················· | ···················· | ···················· | ···················· | ···················· | ···················· |
| A | 8617 hCoV-19/France/IDF-3324/2020\|EPI_ISL_421512\|2020-0 | ···················· | ···················· | ···················· | ···················· | ···················· | ···················· |
| A | 8618 hCoV-19/France/IDF-3345/2020\|EPI_ISL_421513\|2020-0 | ···················· | ···················· | ···················· | ···················· | ···················· | ···················· |
| A | 8619 hCoV-19/France/CVL-3365/2020\|EPI_ISL_421514\|2020-0 | ···················· | ···················· | ···················· | ···················· | ···················· | ···················· |
| A | 8620 hCoV-19/Spain/VC-FISABIO-40/2020\|EPI_ISL_421515\|20 | ···················· | ···················· | ···················· | ···················· | ···················· | ···················· |
| A | 8621 hCoV-19/Spain/VC-FISABIO-52/2020\|EPI_ISL_421519\|20 | ···················· | ···················· | ···················· | ···················· | ···················· | ···················· |
| A | 8622 hCoV-19/USA/WY-WYPHL006/2020\|EPI_ISL_421543\|2020-0 | ···················· | ···················· | ···················· | ···················· | ···················· | ···················· |
| A | 8623 hCoV-19/USA/WY-WYPHL007/2020\|EPI_ISL_421544\|2020-0 | ···················· | ···················· | ···················· | ···················· | ···················· | ···················· |
| A | 8624 hCoV-19/USA/WY-WYPHL008/2020\|EPI_ISL_421545\|2020-0 | ···················· | ···················· | ···················· | ···················· | ···················· | ···················· |
| A | 8625 hCoV-19/USA/WY-WYPHL011/2020\|EPI_ISL_421548\|2020-0 | ···················· | ···················· | ···················· | ···················· | ···················· | ···················· |
| A | 8626 hCoV-19/USA/WY-WYPHL012/2020\|EPI_ISL_421549\|2020-0 | ···················· | ···················· | ···················· | ···················· | ···················· | ···················· |
| A | 8627 hCoV-19/USA/WY-WYPHL014/2020\|EPI_ISL_421550\|2020-0 | ···················· | ···················· | ···················· | ···················· | ···················· | ···················· |
| A | 8628 hCoV-19/USA/WY-WYPHL015/2020\|EPI_ISL_421551\|2020-0 | ···················· | ···················· | ···················· | ···················· | ···················· | ···················· |
| A | 8629 hCoV-19/USA/WY-WYPHL018/2020\|EPI_ISL_421552\|2020-0 | ···················· | ···················· | ···················· | ···················· | ···················· | ···················· |
| A | 8630 hCoV-19/USA/UT-035/2020\|EPI_ISL_421560\|2020-03-18 | ···················· | ···················· | ···················· | ···················· | ···················· | ···················· |
| A | 8631 hCoV-19/USA/UT-055/2020\|EPI_ISL_421561\|2020-03-17 | ···················· | ···················· | ···················· | ···················· | ···················· | ···················· |
| A | 8632 hCoV-19/USA/UT-0139/2020\|EPI_ISL_421562\|2020-03-19 | ···················· | ···················· | ···················· | ···················· | ···················· | ···················· |
| A | 8633 hCoV-19/USA/UT-0364/2020\|EPI_ISL_421563\|2020-03-29 | ···················· | ···················· | ···················· | ···················· | ···················· | ···················· |
| A | 8634 hCoV-19/USA/NY-NYUMC81/2020\|EPI_ISL_421582\|2020-03 | ···················· | ···················· | ···················· | ···················· | ···················· | ···················· |
| A | 8635 hCoV-19/USA/NJ-NYUMC83/2020\|EPI_ISL_421584\|2020-03 | ···················· | ···················· | ···················· | ···················· | ···················· | ···················· |
| A | 8636 hCoV-19/USA/NJ-NYUMC84/2020\|EPI_ISL_421585\|2020-03 | ···················· | ···················· | ···················· | ···················· | ···················· | ···················· |
| A | 8637 hCoV-19/USA/NY-NYUMC85/2020\|EPI_ISL_421586\|2020-03 | ···················· | ···················· | ···················· | ···················· | ···················· | ···················· |
| A | 8638 hCoV-19/USA/NY-NYUMC86/2020\|EPI_ISL_421587\|2020-03 | ···················· | ···················· | ···················· | ···················· | ···················· | ···················· |
| A | 8639 hCoV-19/Netherlands/NA_133/2020\|EPI_ISL_422677\|202 | ···················· | ···················· | ···················· | ···················· | ···················· | ···················· |
| A | 8640 hCoV-19/Netherlands/NA_134/2020\|EPI_ISL_422678\|202 | ···················· | ···················· | ···················· | ···················· | ···················· | ···················· |
| A | 8641 hCoV-19/Netherlands/NA_135/2020\|EPI_ISL_422679\|202 | ···················· | ···················· | ···················· | ···················· | ···················· | ···················· |
| A | 8642 hCoV-19/Netherlands/NA_164/2020\|EPI_ISL_422699\|202 | ···················· | ···················· | ···················· | ···················· | ···················· | ···················· |
| A | 8643 hCoV-19/Netherlands/NA_73/2020\|EPI_ISL_422833\|2020 | ···················· | ···················· | ···················· | ···················· | ···················· | ···················· |
| A | 8644 hCoV-19/Netherlands/NA_74/2020\|EPI_ISL_422834\|2020 | ···················· | ···················· | ···················· | ···················· | ···················· | ···················· |
| A | 8645 hCoV-19/Netherlands/NA_75/2020\|EPI_ISL_422835\|2020 | ···················· | ···················· | ···················· | ···················· | ···················· | ···················· |
| A | 8646 hCoV-19/Netherlands/NA_77/2020\|EPI_ISL_422837\|2020 | ···················· | ···················· | ···················· | ···················· | ···················· | ···················· |
| A | 8647 hCoV-19/Netherlands/NA_78/2020\|EPI_ISL_422838\|2020 | ···················· | ···················· | ···················· | ···················· | ···················· | ···················· |
| A | 8648 hCoV-19/Netherlands/NA_79/2020\|EPI_ISL_422839\|2020 | ···················· | ···················· | ···················· | ············A······· | ···················· | ···················· |
| A | 8649 hCoV-19/Netherlands/NA_80/2020\|EPI_ISL_422840\|2020 | ···················· | ···················· | ···················· | ···················· | ···················· | ···················· |
| A | 8650 hCoV-19/Netherlands/NA_82/2020\|EPI_ISL_422842\|2020 | ···················· | ···················· | ···················· | ···················· | ···················· | ···················· |
| A | 8651 hCoV-19/Netherlands/NA_83/2020\|EPI_ISL_422843\|2020 | ···················· | ···················· | ···················· | ···················· | ···················· | ···················· |
| A | 8652 hCoV-19/Netherlands/NA_85/2020\|EPI_ISL_422845\|2020 | ···················· | ···················· | ···················· | ···················· | ···················· | ···················· |
| A | 8653 hCoV-19/Netherlands/NA_87/2020\|EPI_ISL_422847\|2020 | ·T·················· | ···················· | ···················· | ···················· | ···················· | ···················· |
| A | 8654 hCoV-19/Netherlands/NA_88/2020\|EPI_ISL_422848\|2020 | ···················· | ···················· | ···················· | ···················· | ···················· | ···················· |
| A | 8655 hCoV-19/Netherlands/NA_89/2020\|EPI_ISL_422849\|2020 | ···················· | ···················· | ···················· | ···················· | ···················· | ···················· |
| A | 8656 hCoV-19/Netherlands/NA_90/2020\|EPI_ISL_422850\|2020 | ···················· | ···················· | ···················· | ···················· | ···················· | ···················· |
| A | 8657 hCoV-19/Netherlands/NA_91/2020\|EPI_ISL_422851\|2020 | ···················· | ···················· | ···················· | ···················· | ···················· | ···················· |
| A | 8658 hCoV-19/Netherlands/NA_92/2020\|EPI_ISL_422852\|2020 | ···················· | ···················· | ···················· | ···················· | ···················· | ···················· |
| A | 8659 hCoV-19/Netherlands/NA_93/2020\|EPI_ISL_422853\|2020 | ···················· | ···················· | ···················· | ···················· | ···················· | ···················· |
| A | 8660 hCoV-19/Netherlands/NA_94/2020\|EPI_ISL_422854\|2020 | ···················· | ···················· | ···················· | ···················· | ···················· | ···················· |
| A | 8661 hCoV-19/Netherlands/NA_95/2020\|EPI_ISL_422855\|2020 | ···················· | ···················· | ···················· | ···················· | ···················· | ···················· |
| A | 8662 hCoV-19/Netherlands/NA_96/2020\|EPI_ISL_422856\|2020 | ·T·················· | ···················· | ···················· | ···················· | ···················· | ···················· |
| A | 8663 hCoV-19/Netherlands/NA_97/2020\|EPI_ISL_422857\|2020 | ···················· | ···················· | ···················· | ···················· | ···················· | ···················· |
| A | 8664 hCoV-19/Netherlands/NA_99/2020\|EPI_ISL_422858\|2020 | ···················· | ···················· | ···················· | ···················· | ···················· | ···················· |
| A | 8665 hCoV-19/Netherlands/NoordBrabant_100/2020\|EPI_ISL_ | ···················· | ···················· | ···················· | ···················· | ···················· | ···················· |
| A | 8666 hCoV-19/Netherlands/NoordBrabant_101/2020\|EPI_ISL_ | ···················· | ···················· | ···················· | ···················· | ···················· | ···················· |
| A | 8667 hCoV-19/Netherlands/NoordBrabant_102/2020\|EPI_ISL_ | ···················· | ···················· | ···················· | ···················· | ···················· | ···················· |
| A | 8668 hCoV-19/Netherlands/NoordBrabant_103/2020\|EPI_ISL_ | ···················· | ···················· | ···················· | ···················· | ···················· | ···················· |
| A | 8669 hCoV-19/Netherlands/NoordBrabant_104/2020\|EPI_ISL_ | ···················· | ···················· | ···················· | ···················· | ···················· | ···················· |
| A | 8670 hCoV-19/Netherlands/NoordBrabant_105/2020\|EPI_ISL_ | ···················· | ···················· | ···················· | ···················· | ···················· | ···················· |
| A | 8671 hCoV-19/Netherlands/NoordBrabant_69/2020\|EPI_ISL_4 | ···················· | ···················· | ···················· | ···················· | ···················· | ···················· |
| A | 8672 hCoV-19/Netherlands/NoordBrabant_70/2020\|EPI_ISL_4 | ···················· | ···················· | ···················· | ···················· | ···················· | ···················· |
| A | 8673 hCoV-19/Netherlands/NoordBrabant_71/2020\|EPI_ISL_4 | ···················· | ···················· | ···················· | ···················· | ···················· | ···················· |
| A | 8674 hCoV-19/Netherlands/NoordBrabant_72/2020\|EPI_ISL_4 | ···················· | ···················· | ···················· | ···················· | ···················· | ···················· |
| A | 8675 hCoV-19/Netherlands/NoordBrabant_73/2020\|EPI_ISL_4 | ···················· | ···················· | ···················· | ···················· | ···················· | ···················· |
| A | 8676 hCoV-19/Netherlands/NoordBrabant_74/2020\|EPI_ISL_4 | ···················· | ···················· | ···················· | ···················· | ···················· | ···················· |
| A | 8677 hCoV-19/Netherlands/NoordBrabant_75/2020\|EPI_ISL_4 | ···················· | ···················· | ···················· | ···················· | ···················· | ···················· |
| A | 8678 hCoV-19/Netherlands/NoordBrabant_76/2020\|EPI_ISL_4 | ···················· | ···················· | ···················· | ···················· | ···················· | ···················· |
| A | 8679 hCoV-19/Netherlands/NoordBrabant_77/2020\|EPI_ISL_4 | ···················· | ···················· | ···················· | ···················· | ···················· | ···················· |
| A | 8680 hCoV-19/Netherlands/NoordBrabant_79/2020\|EPI_ISL_4 | ···················· | ···················· | ···················· | ···················· | ···················· | ···················· |
| A | 8681 hCoV-19/Netherlands/NoordBrabant_80/2020\|EPI_ISL_4 | ···················· | ···················· | ···················· | ···················· | ···················· | ···················· |
| A | 8682 hCoV-19/Netherlands/NoordBrabant_81/2020\|EPI_ISL_4 | ···················· | ···················· | ···················· | ···················· | ···················· | ···················· |
| A | 8683 hCoV-19/Netherlands/NoordBrabant_82/2020\|EPI_ISL_4 | ···················· | ···················· | ···················· | ···················· | ···················· | ···················· |
| A | 8684 hCoV-19/Netherlands/NoordBrabant_83/2020\|EPI_ISL_4 | ···················· | ···················· | ···················· | ···················· | ···················· | ···················· |
| A | 8685 hCoV-19/Netherlands/NoordBrabant_85/2020\|EPI_ISL_4 | ···················· | ···················· | ···················· | ···················· | ···················· | ···················· |

```
A    8686 hCoV-19/Netherlands/NoordBrabant_86/2020|EPI_ISL_4   ····················  ····················  ····················  ····················  ····················  ····················
A    8687 hCoV-19/Netherlands/NoordBrabant_87/2020|EPI_ISL_4   ····················  ····················  ····················  ····················  ····················  ····················
A    8688 hCoV-19/Netherlands/NoordBrabant_88/2020|EPI_ISL_4   ····················  ····················  ····················  ····················  ····················  ····················
A    8689 hCoV-19/Netherlands/NoordBrabant_89/2020|EPI_ISL_4   ····················  ····················  ····················  ····················  ····················  ····················
A    8690 hCoV-19/Netherlands/NoordBrabant_90/2020|EPI_ISL_4   ····················  ····················  ····················  ····················  ····················  ····················
A    8691 hCoV-19/Netherlands/NoordBrabant_91/2020|EPI_ISL_4   ····················  ····················  ····················  ····················  ····················  ····················
A    8692 hCoV-19/Netherlands/NoordBrabant_92/2020|EPI_ISL_4   ····················  ····················  ····················  ····················  ····················  ····················
A    8693 hCoV-19/Netherlands/NoordBrabant_93/2020|EPI_ISL_4   ····················  ····················  ····················  ····················  ····················  ····················
A    8694 hCoV-19/Netherlands/NoordBrabant_94/2020|EPI_ISL_4   ····················  ····················  ····················  ····················  ····················  ····················
A    8695 hCoV-19/Netherlands/NoordBrabant_95/2020|EPI_ISL_4   ····················  ····················  ····················  ····················  ····················  ····················
A    8696 hCoV-19/Netherlands/NoordBrabant_96/2020|EPI_ISL_4   ····················  ····················  ····················  ····················  ····················  ····················
A    8697 hCoV-19/Netherlands/NoordBrabant_97/2020|EPI_ISL_4   ····················  ····················  ····················  ····················  ····················  ····················
A    8698 hCoV-19/Netherlands/NoordBrabant_98/2020|EPI_ISL_4   ····················  ····················  ····················  ····················  ····················  ····················
A    8699 hCoV-19/Netherlands/NoordBrabant_99/2020|EPI_ISL_4   ····················  ····················  ····················  ····················  ····················  ····················
A    8700 hCoV-19/Netherlands/NoordHolland_31/2020|EPI_ISL_4   ····················  ····················  ····················  ····················  ····················  ····················
A    8701 hCoV-19/Netherlands/NoordHolland_4/2020|EPI_ISL_42   ····················  ····················  ····················  ····················  ····················  ····················
A    8702 hCoV-19/USA/NY-NYUMC287/2020|EPI_ISL_428801|2020-0   ····················  ····················  ····················  ····················  ····················  ····················
A    8703 hCoV-19/USA/NY-NYUMC290/2020|EPI_ISL_428804|2020-0   ····················  ····················  ····················  ····················  ····················  ····················
A    8704 hCoV-19/USA/NY-NYUMC291/2020|EPI_ISL_428805|2020-0   ····················  ····················  ····················  ····················  ····················  ····················
A    8705 hCoV-19/env/Wuhan/IVDC-HBF13-20/2020|EPI_ISL_40851   ····················  ····················  ····················  ····················  ····················  ····················
A    8706 hCoV-19/env/Wuhan/IVDC-HBF13-21/2020|EPI_ISL_40851   ····················  ····················  ····················  ····················  ····················  ····················
A    8707 hCoV-19/Canada/ON-PHL-2445/2020|EPI_ISL_413014|202   ····················  ·A··················  ····················  ····················  ····················  ····················
A    8708 hCoV-19/Canada/ON_VIDO-01/2020|EPI_ISL_413015|2020   ····················  ·A··················  ··T·················  ····················  ····················  ····················
A    8709 hCoV-19/Brazil/SP-02/2020|EPI_ISL_413016|2020-02-2   ····················  ····················  ····················  ····················  ····················  ····················
A    8710 hCoV-19/South Korea/KUMC01/2020|EPI_ISL_413017|202   ····················  ····················  ····················  ····················  ····················  ····················
A    8711 hCoV-19/South Korea/KUMC02/2020|EPI_ISL_413018|202   ····················  ····················  ····················  ····················  ····················  ····················
A    8712 hCoV-19/Switzerland/ZH-UZH-1000477102/2020|EPI_ISL   ····················  ····················  ····················  ····················  ····················  ····················
A    8713 hCoV-19/Switzerland/ZH-UZH-1000477377/2020|EPI_ISL   ····················  ····················  ····················  ····················  ····················  ····················
A    8714 hCoV-19/Switzerland/ZH-UZH-1000477757/2020|EPI_ISL   ····················  ····················  ····················  ····················  ····················  ····················
A    8715 hCoV-19/Switzerland/ZH-UZH-1000477796/2020|EPI_ISL   ····················  ····················  ····················  ····················  ····················  ····················
A    8716 hCoV-19/Switzerland/ZH-UZH-1000477797/2020|EPI_ISL   ····················  ····················  ····················  ····················  ····················  ····················
A    8717 hCoV-19/Switzerland/ZH-UZH-1000477806/2020|EPI_ISL   ····················  ····················  ····················  ····················  ····················  ····················
A    8718 hCoV-19/USA/WA3-UW1/2020|EPI_ISL_413025|2020-02-27   ····················  ····················  ····················  ····················  ····················  ····················
A    8719 hCoV-19/USA/WA-UW15/2020|EPI_ISL_414363|2020-03-04   ····················  ····················  ····················  ····················  ····················  ····················
A    8720 hCoV-19/USA/WA-UW17/2020|EPI_ISL_414365|2020-03-06   ····················  ····················  ····················  ····················  ····················  ····················
A    8721 hCoV-19/Singapore/12/2020|EPI_ISL_414378|2020-02-1   ····················  ····················  ····················  ····················  ····················  ····················
A    8722 hCoV-19/Singapore/13/2020|EPI_ISL_414379|2020-02-1   ····················  ····················  ····················  ····················  ····················  ····················
A    8723 hCoV-19/Singapore/14/2020|EPI_ISL_414380|2020-02-1   ····················  ····················  ····················  ····················  ····················  ····················
A    8724 hCoV-19/USA/WA-UW61/2020|EPI_ISL_415626|2020-03-10   ····················  ····················  ····················  ····················  ····················  ····················
A    8725 hCoV-19/USA/WA-UW62/2020|EPI_ISL_415627|2020-03-09   ····················  ····················  ····················  ····················  ····················  ····················
A    8726 hCoV-19/Scotland/EDB004/2020|EPI_ISL_415629|2020-0   ····················  ····················  ····················  ····················  ····················  ····················
A    8727 hCoV-19/Scotland/CVR07/2020|EPI_ISL_415630|2020-03   ····················  ····················  ····················  ····················  ····················  ····················
A    8728 hCoV-19/Scotland/CVR10/2020|EPI_ISL_415631|2020-03   ····················  ····················  ····················  ····················  ····················  ····················
A    8729 hCoV-19/Scotland/EDB003/2020|EPI_ISL_415640|2020-0   ····················  ····················  ····················  ····················  ····················  ····················
A    8730 hCoV-19/Georgia/Tb-54/2020|EPI_ISL_415641|2020-02-   ····················  ····················  ····················  ····················  ····················  ····················
A    8731 hCoV-19/Georgia/Tb-477/2020|EPI_ISL_415642|2020-03   ····················  ····················  ····················  ····················  ····················  ····················
A    8732 hCoV-19/Georgia/Tb-468/2020|EPI_ISL_415643|2020-03   ····················  ····················  ····················  ····················  ····················  ····················
A    8733 hCoV-19/Georgia/Tb-82/2020|EPI_ISL_415644|2020-02-   ····················  ····················  ····················  ····················  ····················  ····················
A    8734 hCoV-19/Denmark/SSI-104/2020|EPI_ISL_415648|2020-0   ····················  ····················  ····················  ····················  ····················  ····················
A    8735 hCoV-19/France/HDF-2039/2020|EPI_ISL_415649|2020-0   ····················  ····················  ····················  ····················  ····················  ····················
A    8736 hCoV-19/France/IDF-2075/2020|EPI_ISL_415650|2020-0   ····················  ····················  ····················  ····················  ····················  ····················
A    8737 hCoV-19/France/BFC-2094/2020|EPI_ISL_415651|2020-0   ····················  ····················  ····················  ····················  ····················  ····················
A    8738 hCoV-19/France/BFC-2147/2020|EPI_ISL_415652|2020-0   ····················  ····················  ····················  ····················  ····················  ····················
A    8739 hCoV-19/France/HDF-2174/2020|EPI_ISL_415654|2020-0   ····················  ····················  ····················  ····················  ····················  ····················
A    8740 hCoV-19/Wales/PHW27/2020|EPI_ISL_415655|2020-03-12   ····················  ····················  ····················  ····················  ····················  ····················
A    8741 hCoV-19/Wales/PHW33/2020|EPI_ISL_415656|2020-03-12   ····················  ····················  ····················  ····················  ····················  ····················
A    8742 hCoV-19/Wales/PHW37/2020|EPI_ISL_415657|2020-03-12   ····················  ····A···············  ····················  ····················  ····················  ····················
A    8743 hCoV-19/Chile/RM-MSHS-op2d1/2020|EPI_ISL_415658|20   ····················  ····················  ····················  ····················  ····················  ····················
A    8744 hCoV-19/Wales/PHW28/2020|EPI_ISL_415659|2020-03-12   ····················  ····················  ····················  ····················  ····················  ····················
A    8745 hCoV-19/Chile/RM-MSHS-op3d1/2020|EPI_ISL_415660|20   ····················  ····················  ····················  ····················  ····················  ····················
A    8746 hCoV-19/Chile/RM-MSHS-op4d1/2020|EPI_ISL_415661|20   ····················  ····················  ····················  ····················  ····················  ····················
A    8747 hCoV-19/Spain/CL-ISCIII-201437/2020|EPI_ISL_416994   ····················  ····················  ····················  ····················  ····················  ····················
A    8749 hCoV-19/Hong Kong/HKPU89-2502/2020|EPI_ISL_419250|   ····················  ····················  ····················  ····················  ····················  ····················
A    8750 hCoV-19/Hong Kong/HKPU91-2002/2020|EPI_ISL_419252|   ····················  ····················  ····················  ····················  ····················  ····················
A    8751 hCoV-19/Italy/LAZ-INMI-6/2020|EPI_ISL_419254|2020-   ····················  ····················  ····················  ····················  ····················  ····················
A    8752 hCoV-19/Italy/LAZ-INMI-7/2020|EPI_ISL_419255|2020-   ····················  ····················  ····················  ····················  ····················  ····················
A    8753 hCoV-19/USA/VA-DCLS-0005/2020|EPI_ISL_419256|2020-   ····················  ····················  ····················  ····················  ····················  ····················
A    8754 hCoV-19/USA/VA-DCLS-0006/2020|EPI_ISL_419257|2020-   ····················  ····················  ····················  ····················  ····················  ····················
A    8755 hCoV-19/USA/VA-DCLS-0007/2020|EPI_ISL_419258|2020-   ····················  ····················  ····················  ····················  ····················  ····················
A    8756 hCoV-19/Belgium/UGent-11/2020|EPI_ISL_419259|2020-   ····················  ····················  ····················  ····················  ····················  ····················
A    8757 hCoV-19/USA/VA-DCLS-0008/2020|EPI_ISL_419260|2020-   ····················  ····················  ····················  ····················  ····················  ····················
A    8758 hCoV-19/USA/VA-DCLS-0009/2020|EPI_ISL_419261|2020-   ····················  ····················  ····················  ····················  ····················  ····················
A    8759 hCoV-19/Belgium/UGent-4/2020|EPI_ISL_419266|2020-0   ····················  ····················  ····················  ····················  ····················  ····················
A    8760 hCoV-19/England/SHEF-C053A/2020|EPI_ISL_420235|202   ····················  ····················  ····················  ····················  ····················  ····················
A    8761 hCoV-19/England/SHEF-C0664/2020|EPI_ISL_420236|202   ····················  ····················  ····················  ····················  ····················  ····················
A    8762 hCoV-19/England/SHEF-C0628/2020|EPI_ISL_420237|202   ····················  ····················  ····················  ····················  ····················  ····················
A    8763 hCoV-19/England/SHEF-C051C/2020|EPI_ISL_420238|202   ····················  ····················  ····················  ····················  ····················  ····················
A    8764 hCoV-19/England/SHEF-C0770/2020|EPI_ISL_420239|202   ····················  ····················  ····················  ····················  ····················  ····················
```

| | | | | | | | | |
|---|---|---|---|---|---|---|---|---|
| A | 8765 | hCoV-19/England/SHEF-C0084/2020\|EPI_ISL_420240\|202 | ···················· | ···················· | ···················· | ···················· | ···················· | ···················· |
| A | 8766 | hCoV-19/England/SHEF-C0637/2020\|EPI_ISL_420241\|202 | ···················· | ···················· | ···················· | ···················· | ···················· | ···················· |
| A | 8767 | hCoV-19/England/SHEF-C0734/2020\|EPI_ISL_420242\|202 | ···················· | ···················· | ···················· | ···················· | ···················· | ···················· |
| A | 8768 | hCoV-19/England/SHEF-C05FE/2020\|EPI_ISL_420243\|202 | ···················· | ···················· | ···················· | ···················· | ···················· | ···················· |
| A | 8769 | hCoV-19/England/SHEF-C05B2/2020\|EPI_ISL_420244\|202 | ···················· | ···················· | ···················· | ···················· | ···················· | ···················· |
| A | 8770 | hCoV-19/England/SHEF-C06CE/2020\|EPI_ISL_420245\|202 | ···················· | ···················· | ···················· | ···················· | ···················· | ···················· |
| A | 8771 | hCoV-19/England/SHEF-C05D0/2020\|EPI_ISL_420246\|202 | ···················· | ···················· | ···················· | ···················· | ···················· | ···················· |
| A | 8772 | hCoV-19/England/SHEF-C0585/2020\|EPI_ISL_420247\|202 | ···················· | ·············A······ | ···················· | ···················· | ···················· | ···················· |
| A | 8773 | hCoV-19/England/SHEF-C06BF/2020\|EPI_ISL_420248\|202 | ···················· | ···················· | ···················· | ···················· | ···················· | ···················· |
| A | 8774 | hCoV-19/England/SHEF-C0549/2020\|EPI_ISL_420249\|202 | ···················· | ···················· | ···················· | ···················· | ···················· | ···················· |
| A | 8775 | hCoV-19/England/SHEF-C06DD/2020\|EPI_ISL_420250\|202 | ···················· | ···················· | ···················· | ···················· | ···················· | ···················· |
| A | 8776 | hCoV-19/England/SHEF-C0673/2020\|EPI_ISL_420251\|202 | ···················· | ···················· | ···················· | ···················· | ···················· | ···················· |
| A | 8777 | hCoV-19/England/SHEF-C01AF/2020\|EPI_ISL_420252\|202 | ···················· | ···················· | ···················· | ···················· | ···················· | ···················· |
| A | 8778 | hCoV-19/South Africa/KRISP-06/2020\|EPI_ISL_421573\| | ···················· | ···················· | ···················· | ···················· | ···················· | ···················· |
| A | 8779 | hCoV-19/South Africa/KRISP-07/2020\|EPI_ISL_421574\| | ···················· | ···················· | ···················· | ···················· | ···················· | ···················· |
| A | 8780 | hCoV-19/South Africa/KRISP-011/2020\|EPI_ISL_421575 | ···················· | ···················· | ···················· | ···················· | ···················· | ···················· |
| A | 8781 | hCoV-19/South Africa/KRISP-012/2020\|EPI_ISL_421576 | ···················· | ···················· | ···················· | ···················· | ···················· | ···················· |
| A | 8782 | hCoV-19/USA/NY-NYUMC76/2020\|EPI_ISL_421577\|2020-03 | ···················· | ···················· | ···················· | ···················· | ···················· | ···················· |
| A | 8783 | hCoV-19/USA/NY-NYUMC77/2020\|EPI_ISL_421578\|2020-03 | ···················· | ···················· | ···················· | ···················· | ···················· | ···················· |
| A | 8784 | hCoV-19/USA/NY-NYUMC78/2020\|EPI_ISL_421579\|2020-03 | ···················· | ···················· | ···················· | ···················· | ···················· | ···················· |
| A | 8785 | hCoV-19/USA/NY-NYUMC79/2020\|EPI_ISL_421580\|2020-03 | ···················· | ···················· | ···················· | ···················· | ···················· | ···················· |
| A | 8786 | hCoV-19/USA/NY-NYUMC80/2020\|EPI_ISL_421581\|2020-03 | ···················· | ···················· | ···················· | ···················· | ···················· | ···················· |
| A | 8787 | hCoV-19/USA/NY-NYUMC82/2020\|EPI_ISL_421583\|2020-03 | ···················· | ···················· | ···················· | ···················· | ···················· | ···················· |
| A | 8788 | hCoV-19/Netherlands/NA_40/2020\|EPI_ISL_422801\|2020 | ···················· | ···················· | ···················· | ···················· | ···················· | ···················· |
| A | 8789 | hCoV-19/Netherlands/NA_41/2020\|EPI_ISL_422802\|2020 | ···················· | ···················· | ···················· | ···················· | ···················· | ···················· |
| A | 8790 | hCoV-19/Netherlands/NA_42/2020\|EPI_ISL_422803\|2020 | ···················· | ···················· | ···················· | ···················· | ···················· | ···················· |
| A | 8791 | hCoV-19/Netherlands/NA_43/2020\|EPI_ISL_422804\|2020 | ···················· | ···················· | ···················· | ···················· | ···················· | ···················· |
| A | 8792 | hCoV-19/Netherlands/NA_44/2020\|EPI_ISL_422805\|2020 | ···················· | ···················· | ···················· | ···················· | ···················· | ···················· |
| A | 8793 | hCoV-19/Netherlands/NA_46/2020\|EPI_ISL_422807\|2020 | ···················· | ···················· | ···················· | ···················· | ···················· | ···················· |
| A | 8794 | hCoV-19/Netherlands/NA_47/2020\|EPI_ISL_422808\|2020 | ···················· | ···················· | ···················· | ···················· | ···················· | ···················· |
| A | 8795 | hCoV-19/Netherlands/NA_48/2020\|EPI_ISL_422809\|2020 | ···················· | ···················· | ···················· | ···················· | ···················· | ···················· |
| A | 8796 | hCoV-19/Netherlands/NA_49/2020\|EPI_ISL_422810\|2020 | ···················· | ···················· | ···················· | ···················· | ···················· | ···················· |
| A | 8797 | hCoV-19/Netherlands/NA_50/2020\|EPI_ISL_422811\|2020 | ···················· | ···················· | ···················· | ···················· | ···················· | ···················· |
| A | 8798 | hCoV-19/Netherlands/NA_51/2020\|EPI_ISL_422812\|2020 | ···················· | ···················· | ···················· | ···················· | ···················· | ···················· |
| A | 8799 | hCoV-19/Netherlands/NA_52/2020\|EPI_ISL_422813\|2020 | ···················· | ···················· | ···················· | ···················· | ···················· | ···················· |
| A | 8800 | hCoV-19/Netherlands/NA_54/2020\|EPI_ISL_422815\|2020 | ···················· | ···················· | ···················· | ···················· | ···················· | ···················· |
| A | 8801 | hCoV-19/Netherlands/NA_55/2020\|EPI_ISL_422816\|2020 | ···················· | ···················· | ···················· | ···················· | ···················· | ···················· |
| A | 8802 | hCoV-19/Netherlands/NA_56/2020\|EPI_ISL_422817\|2020 | ···················· | ···················· | ···················· | ···················· | ···················· | ···················· |
| A | 8803 | hCoV-19/Netherlands/NA_57/2020\|EPI_ISL_422818\|2020 | ···················· | ···················· | ···················· | ···················· | ···················· | ···················· |
| A | 8804 | hCoV-19/Netherlands/NA_58/2020\|EPI_ISL_422819\|2020 | ···················· | ···················· | ···················· | ···················· | ···················· | ···················· |
| A | 8805 | hCoV-19/Netherlands/NA_59/2020\|EPI_ISL_422820\|2020 | ···················· | ···················· | ···················· | ···················· | ···················· | ···················· |
| A | 8806 | hCoV-19/Netherlands/NA_60/2020\|EPI_ISL_422821\|2020 | ···················· | ···················· | ···················· | ···················· | ···················· | ···················· |
| A | 8807 | hCoV-19/Netherlands/NA_61/2020\|EPI_ISL_422822\|2020 | ···················· | ···················· | ···················· | ···················· | ···················· | ···················· |
| A | 8808 | hCoV-19/Netherlands/NA_62/2020\|EPI_ISL_422823\|2020 | ···················· | ···················· | ···················· | ···················· | ···················· | ···················· |
| A | 8809 | hCoV-19/Netherlands/NA_64/2020\|EPI_ISL_422825\|2020 | ···················· | ···················· | ···················· | ···················· | ···················· | ···················· |
| A | 8810 | hCoV-19/Netherlands/NA_67/2020\|EPI_ISL_422827\|2020 | ···················· | ···················· | ···················· | ···················· | ···················· | ···················· |
| A | 8811 | hCoV-19/Netherlands/Utrecht_20/2020\|EPI_ISL_422897 | ···················· | ···················· | ···················· | ···················· | ···················· | ···················· |
| A | 8812 | hCoV-19/Netherlands/Utrecht_21/2020\|EPI_ISL_422898 | ···················· | ···················· | ···················· | ···················· | ···················· | ···················· |
| A | 8813 | hCoV-19/Belgium/UGent-1/2020\|EPI_ISL_425048\|2020-0 | ···················· | ···················· | ···················· | ···················· | ···················· | ···················· |
| A | 8814 | hCoV-19/Belgium/UGent-2/2020\|EPI_ISL_425050\|2020-0 | ···················· | ···················· | ···················· | ···················· | ···················· | ···················· |
| A | 8815 | hCoV-19/Belgium/UGent-8/2020\|EPI_ISL_425052\|2020-0 | ···················· | ···················· | ···················· | ···················· | ···················· | ···················· |
| A | 8816 | hCoV-19/Belgium/UGent-9/2020\|EPI_ISL_425053\|2020-0 | ···················· | ···················· | ···················· | ···················· | ···················· | ···················· |
| A | 8817 | hCoV-19/Belgium/UGent-10/2020\|EPI_ISL_425054\|2020- | ···················· | ···················· | ···················· | ···················· | ···················· | ···················· |
| A | 8818 | hCoV-19/Belgium/UGent-13/2020\|EPI_ISL_425055\|2020- | ···················· | ···················· | ···················· | ···················· | ···················· | ···················· |
| A | 8819 | hCoV-19/Belgium/UGent-15/2020\|EPI_ISL_425056\|2020- | ···················· | ···················· | ···················· | ···················· | ···················· | ···················· |
| A | 8820 | hCoV-19/Belgium/UGent-17/2020\|EPI_ISL_425057\|2020- | ···················· | ···················· | ···················· | ···················· | ···················· | ···················· |
| A | 8821 | hCoV-19/Belgium/UGent-18/2020\|EPI_ISL_425058\|2020- | ···················· | ···················· | ···················· | ···················· | ···················· | ···················· |
| A | 8822 | hCoV-19/Belgium/UGent-19/2020\|EPI_ISL_425059\|2020- | ···················· | ···················· | ···················· | ···················· | ···················· | ···················· |
| A | 8823 | hCoV-19/Belgium/UGent-20/2020\|EPI_ISL_425060\|2020- | ···················· | ···················· | ···················· | ···················· | ···················· | ···················· |
| A | 8824 | hCoV-19/Belgium/UGent-21/2020\|EPI_ISL_425061\|2020- | ···················· | ···················· | ···················· | ···················· | ···················· | ···················· |
| A | 8825 | hCoV-19/Belgium/UGent-22/2020\|EPI_ISL_425062\|2020- | ···················· | ···················· | ···················· | ···················· | ···················· | ···················· |
| A | 8826 | hCoV-19/Belgium/UGent-24/2020\|EPI_ISL_425063\|2020- | ···················· | ···················· | ···················· | ···················· | ···················· | ···················· |
| A | 8827 | hCoV-19/Belgium/UGent-25/2020\|EPI_ISL_425064\|2020- | ···················· | ···················· | ···················· | ···················· | ···················· | ···················· |
| A | 8828 | hCoV-19/USA/NY-Wadsworth-26088-01/2020\|EPI_ISL_426 | ···················· | ···················· | ···················· | ···················· | ···················· | ···················· |
| A | 8829 | hCoV-19/USA/NY-Wadsworth-31302-01/2020\|EPI_ISL_426 | ···················· | ···················· | ···················· | ···················· | ···················· | ···················· |
| A | 8830 | hCoV-19/USA/NY-Wadsworth-26856-01/2020\|EPI_ISL_426 | ···················· | ···················· | ···················· | ···················· | ···················· | ···················· |
| A | 8831 | hCoV-19/USA/NY-Wadsworth-31364-01/2020\|EPI_ISL_426 | ···················· | ···················· | ···················· | ···················· | ···················· | ···················· |
| A | 8832 | hCoV-19/USA/NY-Wadsworth-27162-01/2020\|EPI_ISL_426 | ···················· | ···················· | ···················· | ···················· | ···················· | ···················· |
| A | 8833 | hCoV-19/USA/NY-Wadsworth-31930-01/2020\|EPI_ISL_426 | ···················· | ···················· | ···················· | ···················· | ···················· | ···················· |
| A | 8834 | hCoV-19/USA/NY-Wadsworth-16004-01/2020\|EPI_ISL_426 | ···················· | ···················· | ···················· | ···················· | ···················· | ···················· |
| A | 8835 | hCoV-19/USA/NY-Wadsworth-30164-01/2020\|EPI_ISL_426 | ···················· | ···················· | ···················· | ···················· | ···················· | ···················· |
| A | 8836 | hCoV-19/USA/NY-Wadsworth-30246-01/2020\|EPI_ISL_426 | ···················· | ···················· | ···················· | ···················· | ···················· | ···················· |
| A | 8837 | hCoV-19/USA/NY-Wadsworth-32104-01/2020\|EPI_ISL_426 | ···················· | ···················· | ···················· | ···················· | ···················· | ···················· |
| A | 8838 | hCoV-19/USA/NY-Wadsworth-15616-01/2020\|EPI_ISL_426 | ···················· | ···················· | ···················· | ···················· | ···················· | ···················· |
| A | 8839 | hCoV-19/USA/NY-Wadsworth-11736-02/2020\|EPI_ISL_426 | ···················· | ···················· | ···················· | ···················· | ···················· | ···················· |
| A | 8840 | hCoV-19/USA/NY-Wadsworth-30878-01/2020\|EPI_ISL_426 | ···················· | ···················· | ···················· | ···················· | ···················· | ···················· |
| A | 8841 | hCoV-19/USA/NY-Wadsworth-22503-01/2020\|EPI_ISL_426 | ···················· | ···················· | ···················· | ···················· | ···················· | ···················· |
| A | 8842 | hCoV-19/USA/NY-Wadsworth-29275-01/2020\|EPI_ISL_426 | ···················· | ···················· | ···················· | ···················· | ···················· | ···················· |

A 8843 hCoV-19/USA/NY-Wadsworth-22874-01/2020|EPI_ISL_426 ···················· ···················· ····················· ···················· ···················· ····················
A 8844 hCoV-19/USA/NY-Wadsworth-16833-01/2020|EPI_ISL_426 ···················· ···················· ····················· ···················· ···················· ····················
A 8845 hCoV-19/USA/NY-Wadsworth-16480-01/2020|EPI_ISL_426 ···················· ···················· ····················· ···················· ···················· ····················
A 8846 hCoV-19/USA/NY-Wadsworth-26551-01/2020|EPI_ISL_426 ···················· ···················· ····················· ···················· ···················· ····················
A 8847 hCoV-19/USA/NY-Wadsworth-16166-01/2020|EPI_ISL_426 ···················· ···················· ····················· ···················· ···················· ····················
A 8848 hCoV-19/USA/NY-Wadsworth-25860-01/2020|EPI_ISL_426 ···················· ···················· ····················· ···················· ···················· ····················
A 8849 hCoV-19/USA/NY-Wadsworth-31617-01/2020|EPI_ISL_426 ···················· ···················· ····················· ···················· ···················· ····················
A 8850 hCoV-19/USA/NY-Wadsworth-14726-02/2020|EPI_ISL_426 ···················· ···················· ····················· ···················· ···················· ····················
A 8851 hCoV-19/USA/NY-Wadsworth-16766-01/2020|EPI_ISL_426 ···················· ···················· ····················· ···················· ···················· ····················
A 8852 hCoV-19/USA/NY-Wadsworth-11190-01/2020|EPI_ISL_426 ···················· ···················· ····················· ···················· ···················· ····················
A 8853 hCoV-19/USA/NY-Wadsworth-11191-01/2020|EPI_ISL_426 ···················· ···················· ····················· ···················· ···················· ····················
A 8854 hCoV-19/USA/NY-Wadsworth-11245-01/2020|EPI_ISL_426 ···················· ···················· ····················· ···················· ···················· ····················
A 8855 hCoV-19/USA/NY-Wadsworth-11249-01/2020|EPI_ISL_426 ···················· ···················· ····················· ···················· ···················· ····················
A 8856 hCoV-19/USA/NY-Wadsworth-11330-01/2020|EPI_ISL_426 ···················· ···················· ····················· ···················· ···················· ····················
A 8857 hCoV-19/Czech Republic/Seq3/2020|EPI_ISL_426356|20 ···················· ···················· ····················· ···················· ···················· ····················
A 8858 hCoV-19/Czech Republic/Seq4/2020|EPI_ISL_426357|20 ···················· ···················· ····················· ···················· ···················· ····················
A 8859 hCoV-19/Czech Republic/Seq5/2020|EPI_ISL_426358|20 ···················· ···················· ····················· ···················· ···················· ···············A····
A 8860 hCoV-19/Czech Republic/Seq6/2020|EPI_ISL_426359|20 ···················· ···················· ····················· ···················· ···················· ····················
A 8861 hCoV-19/Czech Republic/Seq7/2020|EPI_ISL_426360|20 ···················· ···················· ····················· ···················· ···················· ····················
A 8862 hCoV-19/Mexico/CMX-INCMNSZ-01/2020|EPI_ISL_426361| ···················· ···················· ····················· ···················· ···················· ····················
A 8863 hCoV-19/Mexico/CMX-INCMNSZ-02/2020|EPI_ISL_426362| ···················· ···················· ····················· ···················· ···················· ····················
A 8864 hCoV-19/Mexico/CMX-INCMNSZ-03/2020|EPI_ISL_426363| ···················· ···················· ····················· ···················· ···················· ····················
A 8865 hCoV-19/Mexico/CMX-INCMNSZ-04/2020|EPI_ISL_426364| ···················· ···················· ····················· ···················· ···················· ····················
A 8866 hCoV-19/Mexico/CMX-INCMNSZ-05/2020|EPI_ISL_426365| ···················· ···················· ····················· ···················· ···················· ····················
A 8867 hCoV-19/Slovenia/MB0329/2020|EPI_ISL_426379|2020-0 ···················· ···················· ····················· ···················· ···················· ····················
A 8868 hCoV-19/USA/NY-WCMP12D09P/2020|EPI_ISL_427605|2020 ···················· ···················· ····················· ···················· ···················· ····················
A 8869 hCoV-19/USA/NY-WCMP12E01P/2020|EPI_ISL_427606|2020 ···················· ···················· ····················· ···················· ···················· ····················
A 8870 hCoV-19/USA/NY-WCMP12E06P/2020|EPI_ISL_427607|2020 ···················· ···················· ····················· ···················· ···················· ····················
A 8871 hCoV-19/USA/NY-WCMP12E09P/2020|EPI_ISL_427608|2020 ···················· ···················· ····················· ···················· ···················· ····················
A 8872 hCoV-19/USA/NY-WCMP12F01P/2020|EPI_ISL_427609|2020 ···················· ···················· ····················· ···················· ···················· ····················
A 8873 hCoV-19/USA/NY-WCMP12F02P/2020|EPI_ISL_427610|2020 ···················· ···················· ····················· ···················· ···················· ····················
A 8874 hCoV-19/USA/NY-WCMP12F03P/2020|EPI_ISL_427611|2020 ···················· ···················· ····················· ···················· ···················· ····················
A 8875 hCoV-19/USA/NY-WCMP12F07P/2020|EPI_ISL_427612|2020 ···················· ···················· ····················· ···················· ···················· ····················
A 8876 hCoV-19/USA/NY-WCMP12G01P/2020|EPI_ISL_427613|2020 ···················· ···················· ····················· ···················· ···················· ····················
A 8877 hCoV-19/USA/NY-WCMP12G02P/2020|EPI_ISL_427614|2020 ···················· ···················· ····················· ···················· ···················· ····················
A 8878 hCoV-19/USA/NY-WCMP12G03P/2020|EPI_ISL_427615|2020 ···········T········ ···················· ····················· ···················· ···················· ····················
A 8879 hCoV-19/USA/NY-WCMP12G06P/2020|EPI_ISL_427616|2020 ···················· ···················· ····················· ···················· ···················· ····················
A 8880 hCoV-19/USA/NY-WCMP12G08P/2020|EPI_ISL_427617|2020 ···················· ···················· ····················· ···················· ···················· ····················
A 8881 hCoV-19/USA/NY-WCMP12H01P/2020|EPI_ISL_427618|2020 ···················· ···················· ····················· ···················· ···················· ····················
A 8882 hCoV-19/USA/AK-PHL092/2020|EPI_ISL_427619|2020-04- ···················· ···················· ····················· ···················· ···················· ····················
A 8883 hCoV-19/USA/AK-PHL101/2020|EPI_ISL_427620|2020-04- ···················· ···················· ····················· ···················· ···················· ····················
A 8884 hCoV-19/USA/AK-PHL115/2020|EPI_ISL_427621|2020-03- ···················· ···················· ····················· ···················· ···················· ····················
A 8885 hCoV-19/USA/AK-PHL118/2020|EPI_ISL_427622|2020-03- ···················· ···················· ····················· ···················· ···················· ····················
A 8886 hCoV-19/USA/NY-NYUMC227/2020|EPI_ISL_427627|2020-0 ···················· ···················· ····················· ···················· ···················· ····················
A 8887 hCoV-19/USA/NY-NYUMC228/2020|EPI_ISL_427628|2020-0 ···················· ···················· ····················· ···················· ···················· ····················
A 8888 hCoV-19/USA/NY-NYUMC229/2020|EPI_ISL_427629|2020-0 ···················· ···················· ····················· ···················· ···················· ····················
A 8889 hCoV-19/USA/NY-NYUMC230/2020|EPI_ISL_427630|2020-0 ···················· ···················· ····················· ···················· ···················· ····················
A 8890 hCoV-19/USA/NY-NYUMC231/2020|EPI_ISL_427631|2020-0 ···················· ···················· ····················· ···················· ···················· ····················
A 8891 hCoV-19/USA/NY-NYUMC232/2020|EPI_ISL_427632|2020-0 ···················· ···················· ····················· ···················· ···················· ····················
A 8892 hCoV-19/USA/NY-NYUMC233/2020|EPI_ISL_427633|2020-0 ···················· ···················· ····················· ···················· ···················· ····················
A 8893 hCoV-19/USA/NY-NYUMC235/2020|EPI_ISL_427634|2020-0 ···················· ···················· ····················· ···················· ···················· ····················
A 8894 hCoV-19/USA/NY-NYUMC236/2020|EPI_ISL_427635|2020-0 ···················· ···················· ····················· ···················· ···················· ····················
A 8895 hCoV-19/USA/NY-NYUMC237/2020|EPI_ISL_427636|2020-0 ···················· ···················· ····················· ···················· ···················· ····················
A 8896 hCoV-19/USA/NY-NYUMC238/2020|EPI_ISL_427637|2020-0 ···················· ···················· ····················· ···················· ···················· ····················
A 8897 hCoV-19/USA/NY-NYUMC239/2020|EPI_ISL_427638|2020-0 ···················· ···················· ····················· ···················· ···················· ····················
A 8898 hCoV-19/USA/NY-NYUMC240/2020|EPI_ISL_427639|2020-0 ···················· ···················· ····················· ···················· ···················· ····················
A 8899 hCoV-19/USA/NY-NYUMC241/2020|EPI_ISL_427640|2020-0 ···················· ···················· ····················· ···················· ···················· ····················
A 8900 hCoV-19/USA/NY-NYUMC242/2020|EPI_ISL_427641|2020-0 ···················· ···················· ····················· ···················· ···················· ····················
A 8901 hCoV-19/Australia/NSW219/2020|EPI_ISL_427643|2020- ···················· ···················· ····················· ···················· ···················· ····················
A 8902 hCoV-19/Australia/NSW217/2020|EPI_ISL_427644|2020- ···················· ···················· ····················· ···················· ···················· ····················
A 8903 hCoV-19/Australia/NSW65/2020|EPI_ISL_427645|2020-0 ···················· ···················· ····················· ···················· ···················· ····················
A 8904 hCoV-19/Australia/NSW66/2020|EPI_ISL_427646|2020-0 ···················· ···················· ····················· ···················· ···················· ····················
A 8905 hCoV-19/Australia/NSW155/2020|EPI_ISL_427647|2020- ···················· ···················· ····················· ···················· ···················· ····················
A 8906 hCoV-19/Australia/NSW152/2020|EPI_ISL_427648|2020- ···················· ···················· ····················· ···················· ···················· ····················
A 8907 hCoV-19/Australia/NSW22/2020|EPI_ISL_427649|2020-0 ···················· ···················· ····················· ···················· ···················· ····················
A 8908 hCoV-19/Australia/NSW23/2020|EPI_ISL_427650|2020-0 ···················· ···················· ····················· ···················· ···················· ····················
A 8909 hCoV-19/Australia/NSW29/2020|EPI_ISL_427651|2020-0 ···················· ···················· ····················· ···················· ···················· ····················
A 8910 hCoV-19/Australia/NSW154/2020|EPI_ISL_427652|2020- ···················· ···················· ····················· ···················· ···················· ····················
A 8911 hCoV-19/Australia/NSW153/2020|EPI_ISL_427653|2020- ···················· ···················· ····················· ···················· ···················· ····················
A 8912 hCoV-19/Australia/NSW33/2020|EPI_ISL_427654|2020-0 ···················· ···················· ····················· ···················· ···················· ····················
A 8913 hCoV-19/Australia/NSW34/2020|EPI_ISL_427655|2020-0 ···················· ···················· ····················· ···················· ···················· ····················
A 8914 hCoV-19/Australia/NSW102/2020|EPI_ISL_427656|2020- ···················· ···················· ····················· ···················· ···················· ····················
A 8915 hCoV-19/Australia/NSW103/2020|EPI_ISL_427657|2020- ···················· ···················· ····················· ···················· ···················· ····················
A 8916 hCoV-19/Australia/NSW46/2020|EPI_ISL_427658|2020-0 ···················· ···················· ····················· ···················· ···················· ····················
A 8917 hCoV-19/Australia/NSW49/2020|EPI_ISL_427659|2020-0 ···················· ···················· ····················· ···················· ···················· ····················
A 8918 hCoV-19/Australia/NSW178/2020|EPI_ISL_427670|2020- ···················· ···················· ····················· ···················· ···················· ····················
A 8919 hCoV-19/Australia/NSW212/2020|EPI_ISL_427671|2020- ···················· ···················· ····················· ···················· ···················· ····················
A 8920 hCoV-19/Australia/NSW95/2020|EPI_ISL_427672|2020-0 ···················· ···················· ····················· ···················· ···················· ····················

| | | | | | | | |
|---|---|---|---|---|---|---|---|
| A | 8921 hCoV-19/Australia/NSW135/2020\|EPI_ISL_427673\|2020- | .................... | .................... | ..................... | .................... | .................... | .................... |
| A | 8922 hCoV-19/Australia/NSW157/2020\|EPI_ISL_427674\|2020- | .................... | .................... | ..................... | .................... | .................... | .................... |
| A | 8923 hCoV-19/Australia/NSW92/2020\|EPI_ISL_427675\|2020-0 | .................... | .................... | ..................... | .................... | .................... | .................... |
| A | 8924 hCoV-19/Australia/NSW170/2020\|EPI_ISL_427676\|2020- | .................... | .................... | ..................... | .................... | .................... | .................... |
| A | 8925 hCoV-19/Australia/NSW104/2020\|EPI_ISL_427677\|2020- | .................... | .................... | ..................... | .................... | .................... | .................... |
| A | 8926 hCoV-19/Australia/NSW96/2020\|EPI_ISL_427678\|2020-0 | .................... | .................... | ..................... | .................... | .................... | .................... |
| A | 8927 hCoV-19/Australia/NSW120/2020\|EPI_ISL_427679\|2020- | .................... | .................... | ..................... | .................... | .................... | .................... |
| A | 8928 hCoV-19/Australia/NSW90/2020\|EPI_ISL_427680\|2020-0 | .................... | .................... | ..................... | .................... | .................... | .................... |
| A | 8929 hCoV-19/Australia/NSW93/2020\|EPI_ISL_427681\|2020-0 | .................... | .................... | ..................... | .................... | .................... | .................... |
| A | 8930 hCoV-19/Australia/NSW146/2020\|EPI_ISL_427682\|2020- | .................... | .................... | ..................... | .................... | .................... | .................... |
| A | 8931 hCoV-19/Australia/NSW148/2020\|EPI_ISL_427683\|2020- | .................... | .................... | ..................... | .................... | .................... | .................... |
| A | 8932 hCoV-19/Australia/NSW200/2020\|EPI_ISL_427684\|2020- | .................... | .................... | ..................... | .................... | .................... | .................... |
| A | 8933 hCoV-19/Australia/NSW179/2020\|EPI_ISL_427685\|2020- | .................... | .................... | ..................... | .................... | .................... | .................... |
| A | 8934 hCoV-19/Australia/NSW105/2020\|EPI_ISL_427686\|2020- | .................... | .................... | ..................... | .................... | .................... | .................... |
| A | 8935 hCoV-19/Australia/NSW175/2020\|EPI_ISL_427687\|2020- | .................... | .................... | ..................... | .................... | .................... | .................... |
| A | 8936 hCoV-19/Australia/NSW121/2020\|EPI_ISL_427688\|2020- | .................... | .................... | ..................... | .................... | .................... | .................... |
| A | 8937 hCoV-19/Australia/NSW128/2020\|EPI_ISL_427689\|2020- | .................... | .................... | ..................... | .................... | .................... | .................... |
| A | 8938 hCoV-19/Australia/NSW80/2020\|EPI_ISL_427690\|2020-0 | .................... | .................... | ..................... | .................... | .................... | .................... |
| A | 8939 hCoV-19/Australia/NSW187/2020\|EPI_ISL_427691\|2020- | .................... | .................... | ..................... | .................... | .................... | .................... |
| A | 8940 hCoV-19/Australia/NSW136/2020\|EPI_ISL_427692\|2020- | .................... | .................... | ..................... | .................... | .................... | .................... |
| A | 8941 hCoV-19/Australia/NSW118/2020\|EPI_ISL_427693\|2020- | .................... | .................... | ..................... | .................... | .................... | .................... |
| A | 8942 hCoV-19/Australia/NSW188/2020\|EPI_ISL_427694\|2020- | .................... | .................... | ..................... | .................... | .................... | .................... |
| A | 8943 hCoV-19/Australia/NSW206/2020\|EPI_ISL_427695\|2020- | .................... | .................... | ..................... | .................... | .................... | .................... |
| A | 8944 hCoV-19/Australia/NSW115/2020\|EPI_ISL_427696\|2020- | .................... | .................... | ..................... | .................... | .................... | .................... |
| A | 8945 hCoV-19/Australia/NSW167/2020\|EPI_ISL_427697\|2020- | .................... | .................... | ..................... | .................... | .................... | .................... |
| A | 8946 hCoV-19/Australia/NSW116/2020\|EPI_ISL_427698\|2020- | .................... | .................... | ..................... | .................... | .................... | .................... |
| A | 8947 hCoV-19/Australia/NSW211/2020\|EPI_ISL_427699\|2020- | .................... | .................... | ..................... | .................... | .................... | .................... |
| A | 8948 hCoV-19/USA/WI-UW-260/2020\|EPI_ISL_428936\|2020-04- | .................... | .................... | ..................... | .................... | .................... | .................... |
| A | 8949 hCoV-19/Luxembourg/LNS3259987/2020\|EPI_ISL_428939\| | .................... | .................... | ..................... | .................... | .................... | .................... |
| A | 8950 hCoV-19/Luxembourg/LNS7841004/2020\|EPI_ISL_428940\| | .................... | .................... | ..................... | .................... | .................... | .................... |
| A | 8951 hCoV-19/Luxembourg/LNS6969569/2020\|EPI_ISL_428943\| | .................... | .................... | ..................... | .................... | .................... | .................... |
| A | 8952 hCoV-19/Luxembourg/LNS1387711/2020\|EPI_ISL_428944\| | .................... | .................... | ..................... | .................... | .................... | .................... |
| A | 8953 hCoV-19/Luxembourg/LNS3702558/2020\|EPI_ISL_428945\| | .................... | .................... | ..................... | .................... | .................... | .................... |
| A | 8954 hCoV-19/Luxembourg/LNS0078676/2020\|EPI_ISL_428949\| | .................... | .................... | ..................... | .................... | .................... | .................... |
| A | 8955 hCoV-19/Luxembourg/LNS5971805/2020\|EPI_ISL_428954\| | .................... | .................... | ..................... | .................... | .................... | .................... |
| A | 8956 hCoV-19/Luxembourg/LNS6854244/2020\|EPI_ISL_428956\| | .................... | .................... | ..................... | .................... | .................... | .................... |
| A | 8957 hCoV-19/Luxembourg/LNS4295690/2020\|EPI_ISL_428959\| | .................... | .................... | ..................... | .................... | .................... | .................... |
| A | 8958 hCoV-19/Luxembourg/LNS6326231/2020\|EPI_ISL_428960\| | .................... | .................... | ..................... | .................... | .................... | .................... |
| A | 8959 hCoV-19/Luxembourg/LNS9322962/2020\|EPI_ISL_428961\| | .................... | .................... | ..................... | .................... | .................... | .................... |
| A | 8960 hCoV-19/Luxembourg/LNS6274573/2020\|EPI_ISL_428962\| | .................... | .................... | ..................... | .................... | .................... | .................... |
| A | 8961 hCoV-19/South Korea/KCDC03/2020\|EPI_ISL_407193\|202 | .................... | .................... | ..................... | .................... | .................... | .................... |
| A | 8962 hCoV-19/USA/WA-UW16/2020\|EPI_ISL_414364\|2020-03-05 | .................... | .................... | ..................... | .................... | .................... | .................... |
| A | 8963 hCoV-19/USA/WA-UW18/2020\|EPI_ISL_414366\|2020-03-05 | .................... | .................... | ..................... | .................... | .................... | .................... |
| A | 8964 hCoV-19/USA/WA-UW19/2020\|EPI_ISL_414367\|2020-03-05 | .................... | .................... | ..................... | .................... | .................... | .................... |
| A | 8965 hCoV-19/USA/WA-UW20/2020\|EPI_ISL_414368\|2020-03-05 | .................... | .................... | ..................... | .................... | .................... | .................... |
| A | 8966 hCoV-19/USA/WA-UW21/2020\|EPI_ISL_414369\|2020-03-05 | .................... | .................... | ..................... | .................... | .................... | .................... |
| A | 8967 hCoV-19/USA/WA-UW72/2020\|EPI_ISL_415600\|2020-03-09 | .................... | .................... | ..................... | .................... | .................... | .................... |
| A | 8968 hCoV-19/USA/WA-UW73/2020\|EPI_ISL_415601\|2020-03-10 | .................... | .................... | ..................... | .................... | .................... | .................... |
| A | 8969 hCoV-19/USA/WA-UW74/2020\|EPI_ISL_415602\|2020-03-10 | .................... | .................... | ..................... | .................... | .................... | .................... |
| A | 8970 hCoV-19/USA/WA-UW75/2020\|EPI_ISL_415603\|2020-03-10 | .................... | .................... | ..................... | .................... | .................... | .................... |
| A | 8971 hCoV-19/USA/WA-UW76/2020\|EPI_ISL_415604\|2020-03-10 | .................... | .................... | ..................... | .................... | .................... | .................... |
| A | 8972 hCoV-19/USA/WA-UW40/2020\|EPI_ISL_415605\|2020-03-05 | .................... | .................... | ..................... | .................... | .................... | .................... |
| A | 8973 hCoV-19/USA/WA-UW41/2020\|EPI_ISL_415606\|2020-03-08 | .................... | .................... | ..................... | .................... | .................... | .................... |
| A | 8974 hCoV-19/USA/WA-UW42/2020\|EPI_ISL_415607\|2020-03-09 | .................... | .................... | ..................... | .................... | .................... | .................... |
| A | 8975 hCoV-19/USA/WA-UW43/2020\|EPI_ISL_415608\|2020-03-08 | .................... | .................... | ..................... | .................... | .................... | .................... |
| A | 8976 hCoV-19/USA/WA-UW44/2020\|EPI_ISL_415609\|2020-03-08 | .................... | .................... | ..................... | .................... | .................... | .................... |
| A | 8977 hCoV-19/USA/WA-UW45/2020\|EPI_ISL_415610\|2020-03-08 | .................... | .................... | ..................... | .................... | .................... | .................... |
| A | 8978 hCoV-19/USA/WA-UW46/2020\|EPI_ISL_415611\|2020-03-09 | .................... | .................... | ..................... | .................... | .................... | .................... |
| A | 8979 hCoV-19/USA/WA-UW47/2020\|EPI_ISL_415612\|2020-03-09 | .................... | .................... | ..................... | .................... | .................... | .................... |
| A | 8980 hCoV-19/USA/WA-UW48/2020\|EPI_ISL_415613\|2020-03-09 | .................... | .................... | ..................... | .................... | .................... | .................... |
| A | 8981 hCoV-19/USA/WA-UW49/2020\|EPI_ISL_415614\|2020-03-09 | .................... | .................... | ..................... | .................... | .................... | .................... |
| A | 8982 hCoV-19/USA/WA-UW50/2020\|EPI_ISL_415615\|2020-03-08 | .................... | .................... | ..................... | .................... | .................... | .................... |
| A | 8983 hCoV-19/USA/WA-UW51/2020\|EPI_ISL_415616\|2020-03-08 | .................... | .................... | ..................... | .................... | .................... | .................... |
| A | 8984 hCoV-19/USA/WA-UW52/2020\|EPI_ISL_415617\|2020-03-09 | .................... | .................... | ..................... | .................... | .................... | .................... |
| A | 8985 hCoV-19/USA/WA-UW54/2020\|EPI_ISL_415619\|2020-03-09 | .................... | .................... | ..................... | .................... | .................... | .................... |
| A | 8986 hCoV-19/USA/WA-UW55/2020\|EPI_ISL_415620\|2020-03-09 | .................... | .................... | ..................... | .................... | .................... | .................... |
| A | 8987 hCoV-19/USA/WA-UW56/2020\|EPI_ISL_415621\|2020-03-09 | .................... | .................... | ..................... | .................... | .................... | .................... |
| A | 8988 hCoV-19/USA/WA-UW57/2020\|EPI_ISL_415622\|2020-03-09 | .................... | .................... | ..................... | .................... | .................... | .................... |
| A | 8989 hCoV-19/USA/WA-UW59/2020\|EPI_ISL_415624\|2020-03-09 | .................... | .................... | ..................... | .................... | .................... | .................... |
| A | 8990 hCoV-19/USA/WA-UW60/2020\|EPI_ISL_415625\|2020-03-09 | .................... | .................... | ..................... | .................... | .................... | .................... |
| A | 8991 hCoV-19/Switzerland/GR-HUG-2988/2020\|EPI_ISL_41569 | .................... | .................... | ..................... | .................... | .................... | .................... |
| A | 8992 hCoV-19/Switzerland/GR-HUG-3043/2020\|EPI_ISL_41569 | .................... | .................... | ..................... | .................... | .................... | .................... |
| A | 8993 hCoV-19/Malaysia/MKAK-CL-2020-7554/2020\|EPI_ISL_41 | .................... | .................... | ..................... | .................... | .................... | .................... |
| A | 8994 hCoV-19/France/ARA-10552/2020\|EPI_ISL_419168\|2020- | .................... | .................... | ..................... | .................... | .................... | .................... |
| A | 8995 hCoV-19/France/ARA-11939/2020\|EPI_ISL_419169\|2020- | .................... | .................... | ..................... | .................... | .................... | .................... |
| A | 8996 hCoV-19/France/ARA-11943/2020\|EPI_ISL_419170\|2020- | .................... | .................... | ..................... | .................... | .................... | .................... |
| A | 8997 hCoV-19/France/ARA-11949/2020\|EPI_ISL_419171\|2020- | .................... | .................... | ..................... | .................... | .................... | .................... |
| A | 8998 hCoV-19/France/ARA-11950/2020\|EPI_ISL_419172\|2020- | .................... | .................... | ..................... | .................... | .................... | .................... |

| | | | | | | | | | |
|---|---|---|---|---|---|---|---|---|---|
| A | 8999 | hCoV-19/France/ARA-11952/2020\|EPI_ISL_419173\|2020- | .................... | .................... | .................... | .................... | .................... | .................... | |
| A | 9000 | hCoV-19/France/ARA-11980/2020\|EPI_ISL_419174\|2020- | .................... | .................... | .................... | .................... | .................... | .................... | |
| A | 9001 | hCoV-19/France/ARA-11995/2020\|EPI_ISL_419175\|2020- | .................... | .................... | .................... | .................... | .................... | .................... | |
| A | 9002 | hCoV-19/France/ARA-11997/2020\|EPI_ISL_419176\|2020- | .................... | .................... | .................... | .................... | .................... | .................... | |
| A | 9003 | hCoV-19/France/ARA-12125/2020\|EPI_ISL_419177\|2020- | .................... | .................... | .................... | .................... | .................... | .................... | |
| A | 9004 | hCoV-19/France/ARA-12217/2020\|EPI_ISL_419178\|2020- | .................... | .................... | .................... | .................... | .................... | .................... | |
| A | 9005 | hCoV-19/France/ARA-12222/2020\|EPI_ISL_419179\|2020- | .................... | .................... | .................... | .................... | .................... | .................... | |
| A | 9006 | hCoV-19/France/ARA-12238/2020\|EPI_ISL_419180\|2020- | .................... | .................... | .................... | .................... | .................... | .................... | |
| A | 9007 | hCoV-19/France/ARA-12249/2020\|EPI_ISL_419181\|2020- | .................... | .................... | .................... | .................... | .................... | .................... | |
| A | 9008 | hCoV-19/France/ARA-12250/2020\|EPI_ISL_419182\|2020- | .................... | .................... | .................... | .................... | .................... | .................... | |
| A | 9009 | hCoV-19/France/ARA-12253/2020\|EPI_ISL_419183\|2020- | .................... | .................... | .................... | .................... | .................... | .................... | |
| A | 9010 | hCoV-19/France/ARA-12260/2020\|EPI_ISL_419184\|2020- | .................... | .................... | .................... | .................... | .................... | .................... | |
| A | 9011 | hCoV-19/France/ARA-12264/2020\|EPI_ISL_419185\|2020- | .................... | .................... | .................... | .................... | .................... | .................... | |
| A | 9012 | hCoV-19/France/ARA-12265/2020\|EPI_ISL_419186\|2020- | .................... | .................... | .................... | .................... | .................... | .................... | |
| A | 9013 | hCoV-19/France/ARA-12269/2020\|EPI_ISL_419187\|2020- | .................... | .................... | .................... | .................... | .................... | .................... | |
| A | 9014 | hCoV-19/France/ARA-12270/2020\|EPI_ISL_419188\|2020- | .................... | .................... | .................... | .................... | .................... | .................... | |
| A | 9015 | hCoV-19/Norway/1694/2020\|EPI_ISL_420136\|2020-03-04 | .................... | .................... | .................... | .................... | .................... | .................... | |
| A | 9016 | hCoV-19/Norway/1772/2020\|EPI_ISL_420137\|2020-03-08 | .................... | .................... | .................... | .................... | .................... | .................... | |
| A | 9017 | hCoV-19/Norway/1811/2020\|EPI_ISL_420139\|2020-03-05 | .................... | .................... | .................... | .................... | .................... | .................... | |
| A | 9018 | hCoV-19/Georgia/Tb-1352/2020\|EPI_ISL_420140\|2020-0 | .................... | .................... | .................... | .................... | .................... | .................... | |
| A | 9019 | hCoV-19/Norway/1867/2020\|EPI_ISL_420141\|2020-03-10 | .................... | .................... | .................... | .................... | .................... | .................... | |
| A | 9020 | hCoV-19/Georgia/Tb-1523/2020\|EPI_ISL_420142\|2020-0 | .................... | .................... | .................... | .................... | .................... | .................... | |
| A | 9021 | hCoV-19/Norway/1917/2020\|EPI_ISL_420143\|2020-03-10 | .................... | .................... | .................... | .................... | .................... | .................... | |
| A | 9022 | hCoV-19/Georgia/Tb-1679/2020\|EPI_ISL_420144\|2020-0 | .................... | .................... | .................... | .................... | .................... | .................... | |
| A | 9023 | hCoV-19/Norway/1951/2020\|EPI_ISL_420145\|2020-03-09 | .................... | .................... | .................... | .................... | .................... | .................... | |
| A | 9024 | hCoV-19/Norway/1955/2020\|EPI_ISL_420146\|2020-03-11 | .................... | .................... | .................... | .................... | .................... | .................... | |
| A | 9025 | hCoV-19/Norway/1989/2020\|EPI_ISL_420147\|2020-03-10 | .................... | .................... | .................... | .................... | .................... | .................... | |
| A | 9026 | hCoV-19/Norway/2084/2020\|EPI_ISL_420150\|2020-03-09 | .................... | .................... | .................... | .................... | .................... | .................... | |
| A | 9027 | hCoV-19/Norway/2087/2020\|EPI_ISL_420151\|2020-03-17 | .................... | .................... | .................... | .................... | .................... | .................... | |
| A | 9028 | hCoV-19/Norway/2088/2020\|EPI_ISL_420152\|2020-03-17 | .................... | .A.................. | .................... | .................... | .................... | .................... | |
| A | 9031 | hCoV-19/England/SHEF-C07AD/2020\|EPI_ISL_420155\|202 | .................... | .................... | .................... | .................... | .................... | .................... | |
| A | 9032 | hCoV-19/England/SHEF-C07DA/2020\|EPI_ISL_420156\|202 | .................... | .................... | .................... | .................... | .................... | .................... | |
| A | 9033 | hCoV-19/England/SHEF-C079E/2020\|EPI_ISL_420157\|202 | .................... | .................... | .................... | .................... | .................... | .................... | |
| A | 9034 | hCoV-19/England/SHEF-C07CB/2020\|EPI_ISL_420158\|202 | .................... | .................... | .................... | .................... | .................... | .................... | |
| A | 9035 | hCoV-19/England/SHEF-C07BC/2020\|EPI_ISL_420159\|202 | .................... | .................... | .................... | .................... | .................... | .................... | |
| A | 9036 | hCoV-19/England/SHEF-C07E9/2020\|EPI_ISL_420160\|202 | .................... | .................... | .................... | .................... | .................... | .................... | |
| A | 9037 | hCoV-19/England/SHEF-C08E6/2020\|EPI_ISL_420161\|202 | .................... | .................... | .................... | .................... | .................... | .................... | |
| A | 9038 | hCoV-19/England/SHEF-C0831/2020\|EPI_ISL_420162\|202 | .................... | .................... | .................... | .................... | .................... | .................... | |
| A | 9039 | hCoV-19/England/SHEF-C086E/2020\|EPI_ISL_420163\|202 | .................... | .................... | .................... | .................... | .................... | .................... | |
| A | 9040 | hCoV-19/England/SHEF-C08AA/2020\|EPI_ISL_420164\|202 | .................... | .................... | .................... | .................... | .................... | .................... | |
| A | 9041 | hCoV-19/England/SHEF-C08C8/2020\|EPI_ISL_420165\|202 | .................... | .................... | .................... | .................... | .................... | .................... | |
| A | 9042 | hCoV-19/England/SHEF-C0813/2020\|EPI_ISL_420166\|202 | .................... | .................... | .................... | .................... | .................... | .................... | |
| A | 9043 | hCoV-19/England/SHEF-C0804/2020\|EPI_ISL_420167\|202 | .................... | .................... | .................... | .................... | .................... | .................... | |
| A | 9044 | hCoV-19/England/SHEF-C087D/2020\|EPI_ISL_420168\|202 | .................... | .................... | .................... | .................... | .................... | .................... | |
| A | 9045 | hCoV-19/England/SHEF-C0822/2020\|EPI_ISL_420169\|202 | .................... | ...................A | .................... | .................... | .................... | .................... | 1 |
| A | 9046 | hCoV-19/England/SHEF-C0901/2020\|EPI_ISL_420170\|202 | .................... | .................... | .................... | .................... | .................... | .................... | |
| A | 9047 | hCoV-19/England/SHEF-C08B9/2020\|EPI_ISL_420171\|202 | .................... | .................... | .................... | .................... | .................... | .................... | |
| A | 9048 | hCoV-19/England/SHEF-C089B/2020\|EPI_ISL_420172\|202 | .................... | .................... | .................... | .................... | .................... | .................... | |
| A | 9049 | hCoV-19/England/SHEF-C08F5/2020\|EPI_ISL_420173\|202 | .................... | .................... | .................... | .................... | .................... | .................... | |
| A | 9050 | hCoV-19/England/SHEF-C088C/2020\|EPI_ISL_420174\|202 | .................... | .................... | .................... | .................... | .................... | .................... | |
| A | 9051 | hCoV-19/England/SHEF-C07F8/2020\|EPI_ISL_420175\|202 | .................... | .................... | .................... | .................... | .................... | .................... | |
| A | 9053 | hCoV-19/England/SHEF-C0840/2020\|EPI_ISL_420177\|202 | .................... | .................... | .................... | .................... | .................... | .................... | |
| A | 9054 | hCoV-19/England/SHEF-C04A6/2020\|EPI_ISL_420178\|202 | .................... | .................... | .................... | .................... | .................... | .................... | |
| A | 9055 | hCoV-19/England/SHEF-C042E/2020\|EPI_ISL_420179\|202 | .................... | .................... | .................... | .................... | .................... | .................... | |
| A | 9056 | hCoV-19/England/SHEF-C02CA/2020\|EPI_ISL_420180\|202 | .................... | .................... | .................... | .................... | .................... | .................... | |
| A | 9057 | hCoV-19/England/SHEF-C0321/2020\|EPI_ISL_420181\|202 | .................... | .................... | .................... | .................... | .................... | .................... | |
| A | 9058 | hCoV-19/England/SHEF-C029D/2020\|EPI_ISL_420182\|202 | .................... | .................... | .................... | .................... | .................... | .................... | |
| A | 9060 | hCoV-19/England/SHEF-C03C7/2020\|EPI_ISL_420184\|202 | .................... | .................... | .................... | .................... | .................... | .................... | |
| A | 9061 | hCoV-19/England/SHEF-C038B/2020\|EPI_ISL_420185\|202 | .................... | .................... | .................... | .................... | .................... | .................... | |
| A | 9062 | hCoV-19/England/SHEF-C0479/2020\|EPI_ISL_420186\|202 | .................... | .................... | .................... | .................... | .................... | .................... | |
| A | 9064 | hCoV-19/England/SHEF-C0400/2020\|EPI_ISL_420188\|202 | .................... | .................... | .................... | .................... | .................... | .................... | |
| A | 9065 | hCoV-19/England/SHEF-C0497/2020\|EPI_ISL_420189\|202 | .................... | .................... | .................... | .................... | .................... | .................... | |
| A | 9066 | hCoV-19/England/SHEF-C03F4/2020\|EPI_ISL_420190\|202 | .................... | .................... | .................... | .................... | .................... | .................... | |
| A | 9067 | hCoV-19/England/SHEF-C0260/2020\|EPI_ISL_420191\|202 | .................... | .................... | .................... | .................... | ..............N..... | ...N................ | |
| A | 9068 | hCoV-19/England/SHEF-C02BB/2020\|EPI_ISL_420192\|202 | .................... | .................... | .................... | .................... | .................... | .................... | |
| A | 9069 | hCoV-19/England/SHEF-C044C/2020\|EPI_ISL_420193\|202 | .................... | .................... | .................... | .................... | .................... | .................... | |
| A | 9070 | hCoV-19/England/SHEF-C028E/2020\|EPI_ISL_420194\|202 | .................... | .................... | .................... | .................... | .................... | .................... | |
| A | 9071 | hCoV-19/England/SHEF-C039A/2020\|EPI_ISL_420195\|202 | .................... | .................... | .................... | .................... | .................... | .................... | |
| A | 9072 | hCoV-19/England/SHEF-C0251/2020\|EPI_ISL_420196\|202 | .................... | .................... | .................... | .................... | .................... | .................... | |
| A | 9073 | hCoV-19/England/SHEF-C03B8/2020\|EPI_ISL_420197\|202 | .................... | .................... | .................... | .................... | .................... | .................... | |
| A | 9074 | hCoV-19/England/SHEF-C0206/2020\|EPI_ISL_420198\|202 | .................... | .................... | .................... | .................... | .................... | .................... | |
| A | 9075 | hCoV-19/England/SHEF-C0233/2020\|EPI_ISL_420199\|202 | .................... | .................... | .................... | .................... | .................... | .................... | |
| A | 9076 | hCoV-19/Portugal/PT0064/2020\|EPI_ISL_421467\|2020-0 | .................... | .................... | .................... | .................... | .................... | .................... | |
| A | 9077 | hCoV-19/Portugal/PT0065/2020\|EPI_ISL_421468\|2020-0 | .................... | .................... | .................... | .................... | .................... | .................... | |
| A | 9078 | hCoV-19/Portugal/PT0066/2020\|EPI_ISL_421469\|2020-0 | .................... | .................... | .................... | .................... | .................... | .................... | |
| A | 9079 | hCoV-19/Portugal/PT0067/2020\|EPI_ISL_421470\|2020-0 | .................... | .................... | .................... | .................... | .................... | .................... | |
| A | 9080 | hCoV-19/Portugal/PT0068/2020\|EPI_ISL_421471\|2020-0 | .................... | .................... | .................... | .................... | .................... | .................... | |
| A | 9081 | hCoV-19/Portugal/PT0069/2020\|EPI_ISL_421472\|2020-0 | .................... | .................... | .................... | .................... | .................... | .................... | |

| | | | | | | | | |
|---|---|---|---|---|---|---|---|---|
| A | 9082 | hCoV-19/Portugal/PT0070/2020\|EPI_ISL_421473\|2020-0 | .................... | .................... | .................... | .................... | .................... | .................... |
| A | 9083 | hCoV-19/Portugal/PT0071/2020\|EPI_ISL_421474\|2020-0 | .................... | .................... | .................... | .................... | .................... | .................... |
| A | 9084 | hCoV-19/Portugal/PT0072/2020\|EPI_ISL_421475\|2020-0 | .................... | .................... | .................... | .................... | .................... | .................... |
| A | 9085 | hCoV-19/Portugal/PT0073/2020\|EPI_ISL_421476\|2020-0 | .................... | .................... | .................... | .................... | .................... | .................... |
| A | 9086 | hCoV-19/Portugal/PT0074/2020\|EPI_ISL_421477\|2020-0 | .................... | .................... | .................... | .................... | .................... | .................... |
| A | 9087 | hCoV-19/Portugal/PT0075/2020\|EPI_ISL_421478\|2020-0 | .................... | .................... | .................... | .................... | .................... | .................... |
| A | 9088 | hCoV-19/Portugal/PT0076/2020\|EPI_ISL_421479\|2020-0 | .................... | .................... | .................... | .................... | .................... | .................... |
| A | 9089 | hCoV-19/Portugal/PT0077/2020\|EPI_ISL_421480\|2020-0 | .................... | .................... | .................... | .................... | .................... | .................... |
| A | 9090 | hCoV-19/Portugal/PT0078/2020\|EPI_ISL_421481\|2020-0 | .................... | .................... | .................... | .................... | .................... | .................... |
| A | 9091 | hCoV-19/Portugal/PT0079/2020\|EPI_ISL_421482\|2020-0 | .................... | .................... | .................... | .................... | .................... | .................... |
| A | 9092 | hCoV-19/Portugal/PT0080/2020\|EPI_ISL_421483\|2020-0 | .................... | .................... | .................... | .................... | .................... | .................... |
| A | 9093 | hCoV-19/Portugal/PT0081/2020\|EPI_ISL_421484\|2020-0 | .................... | .................... | .................... | .................... | .................... | .................... |
| A | 9094 | hCoV-19/Portugal/PT0082/2020\|EPI_ISL_421485\|2020-0 | .................... | .................... | .................... | .................... | .................... | .................... |
| A | 9095 | hCoV-19/Portugal/PT0083/2020\|EPI_ISL_421486\|2020-0 | .................... | .................... | .................... | .................... | .................... | .................... |
| A | 9096 | hCoV-19/Portugal/PT0084/2020\|EPI_ISL_421487\|2020-0 | .................... | .................... | .................... | .................... | .................... | .................... |
| A | 9097 | hCoV-19/Portugal/PT0085/2020\|EPI_ISL_421488\|2020-0 | .................... | .................... | .................... | .................... | .................... | .................... |
| A | 9098 | hCoV-19/Portugal/PT0086/2020\|EPI_ISL_421489\|2020-0 | .................... | .................... | .................... | .................... | .................... | .................... |
| A | 9099 | hCoV-19/Portugal/PT0087/2020\|EPI_ISL_421490\|2020-0 | .................... | .................... | .................... | .................... | .................... | .................... |
| A | 9100 | hCoV-19/Portugal/PT0088/2020\|EPI_ISL_421491\|2020-0 | .................... | .................... | .................... | .................... | .................... | .................... |
| A | 9101 | hCoV-19/Portugal/PT0089/2020\|EPI_ISL_421492\|2020-0 | .................... | .................... | .................... | .................... | .................... | .................... |
| A | 9102 | hCoV-19/Portugal/PT0091/2020\|EPI_ISL_421494\|2020-0 | .................... | .................... | .................... | .................... | .................... | .................... |
| A | 9103 | hCoV-19/Portugal/PT0092/2020\|EPI_ISL_421495\|2020-0 | .................... | .................... | .................... | .................... | .................... | .................... |
| A | 9104 | hCoV-19/Portugal/PT0093/2020\|EPI_ISL_421496\|2020-0 | .................... | .................... | .................... | .................... | .................... | .................... |
| A | 9105 | hCoV-19/Portugal/PT0094/2020\|EPI_ISL_421497\|2020-0 | .................... | .................... | .................... | .................... | .................... | .................... |
| A | 9106 | hCoV-19/Portugal/PT0095/2020\|EPI_ISL_421498\|2020-0 | .................... | .................... | .................... | .................... | .................... | .................... |
| A | 9107 | hCoV-19/Portugal/PT0096/2020\|EPI_ISL_421499\|2020-0 | .................... | .................... | .................... | .................... | .................... | .................... |
| A | 9108 | hCoV-19/Netherlands/NA_37/2020\|EPI_ISL_422798\|2020 | .................... | .................... | .................... | .................... | .................... | .................... |
| A | 9109 | hCoV-19/Netherlands/NA_38/2020\|EPI_ISL_422799\|2020 | .................... | .................... | .................... | .................... | .................... | .................... |
| A | 9110 | hCoV-19/USA/NY-WCMP12C01P/2020\|EPI_ISL_427600\|2020 | .................... | .................... | .................... | .................... | .................... | .................... |
| A | 9111 | hCoV-19/USA/NY-WCMP12C04P/2020\|EPI_ISL_427601\|2020 | .................... | .................... | .................... | .................... | .................... | .................... |
| A | 9112 | hCoV-19/USA/NY-WCMP12C09P/2020\|EPI_ISL_427602\|2020 | .................... | .................... | .................... | .................... | .................... | .................... |
| A | 9113 | hCoV-19/USA/NY-WCMP12D02P/2020\|EPI_ISL_427603\|2020 | .................... | .................... | .................... | .................... | .................... | .................... |
| A | 9114 | hCoV-19/USA/NY-WCMP12D08P/2020\|EPI_ISL_427604\|2020 | .................... | .................... | .................... | .................... | .................... | .................... |
| A | 9115 | hCoV-19/Australia/NSW51/2020\|EPI_ISL_427660\|2020-0 | .................... | .................... | .................... | .................... | .................... | .................... |
| A | 9116 | hCoV-19/Australia/NSW111/2020\|EPI_ISL_427661\|2020- | .................... | .................... | .................... | .................... | .................... | .................... |
| A | 9117 | hCoV-19/Australia/NSW59/2020\|EPI_ISL_427662\|2020-0 | .................... | .................... | .................... | .................... | .................... | .................... |
| A | 9118 | hCoV-19/Australia/NSW60/2020\|EPI_ISL_427663\|2020-0 | .................... | .................... | .................... | .................... | .................... | .................... |
| A | 9119 | hCoV-19/Australia/NSW63/2020\|EPI_ISL_427664\|2020-0 | .................... | .................... | .................... | .................... | .................... | .................... |
| A | 9120 | hCoV-19/Australia/NSW218/2020\|EPI_ISL_427665\|2020- | .................... | .................... | .................... | .................... | .................... | .................... |
| A | 9121 | hCoV-19/Australia/NSW112/2020\|EPI_ISL_427666\|2020- | .................... | .................... | .................... | .................... | .................... | .................... |
| A | 9122 | hCoV-19/Australia/NSW177/2020\|EPI_ISL_427667\|2020- | .................... | .................... | .................... | .................... | .................... | .................... |
| A | 9123 | hCoV-19/Australia/NSW174/2020\|EPI_ISL_427668\|2020- | .................... | .................... | .................... | .................... | .................... | .................... |
| A | 9124 | hCoV-19/Australia/NSW119/2020\|EPI_ISL_427669\|2020- | .................... | .................... | .................... | .................... | .................... | .................... |
| A | 9125 | hCoV-19/Russia/Moscow-80112/2020\|EPI_ISL_428900\|20 | .................... | .................... | .................... | .................... | .................... | .................... |
| A | 9126 | hCoV-19/Crimea/SRC-80603/2020\|EPI_ISL_428901\|2020- | .................... | .................... | .................... | .................... | .................... | .................... |
| A | 9127 | hCoV-19/Russia/Orenburg-80305/2020\|EPI_ISL_428902\| | .................... | .................... | .................... | .................... | .................... | .................... |
| A | 9128 | hCoV-19/Russia/Primorskiy-79001/2020\|EPI_ISL_42890 | .................... | .................... | .................... | .................... | .................... | .................... |
| A | 9129 | hCoV-19/Russia/Vector_77628/2020\|EPI_ISL_428904\|20 | .................... | .................... | .................... | .................... | .................... | .................... |
| A | 9130 | hCoV-19/Russia/Moscow-80402/2020\|EPI_ISL_428905\|20 | .................... | .................... | .................... | .................... | .................... | .................... |
| A | 9131 | hCoV-19/Russia/Buryatia-84504/2020\|EPI_ISL_428906\| | .................... | .................... | .................... | .................... | .................... | .................... |
| A | 9132 | hCoV-19/Russia/Sakha-89702/2020\|EPI_ISL_428907\|202 | .................... | .................... | .................... | .................... | .................... | .................... |
| A | 9133 | hCoV-19/Russia/Pskov-83602/2020\|EPI_ISL_428908\|202 | .................... | .................... | .................... | .................... | .................... | .................... |
| A | 9134 | hCoV-19/Russia/Chechenya-83801/2020\|EPI_ISL_428909 | .................... | .................... | .................... | .................... | .................... | .................... |
| A | 9135 | hCoV-19/Russia/Khabarovsk-88201/2020\|EPI_ISL_42891 | .................... | .................... | .................... | .................... | .................... | .................... |
| A | 9136 | hCoV-19/Russia/Buryatia-88903/2020\|EPI_ISL_428911\| | .................... | .................... | .................... | .................... | .................... | .................... |
| A | 9137 | hCoV-19/Russia/Sakhalin-84401/2020\|EPI_ISL_428912\| | .................... | .................... | .................... | .................... | .................... | .................... |
| A | 9138 | hCoV-19/Russia/Buryatia-84508/2020\|EPI_ISL_428913\| | .................... | .................... | .................... | .................... | .................... | .................... |
| A | 9139 | hCoV-19/Russia/Buryatia-87104/2020\|EPI_ISL_428914\| | .................... | .................... | .................... | .................... | .................... | .................... |
| A | 9140 | hCoV-19/Russia/Sverdlovsk-82904/2020\|EPI_ISL_42891 | .................... | .................... | .................... | .................... | .................... | .................... |
| A | 9141 | hCoV-19/Russia/Chechenya-83803/2020\|EPI_ISL_428916 | .................... | .................... | .................... | .................... | .................... | .................... |
| A | 9142 | hCoV-19/Russia/Tatarstan-82208/2020\|EPI_ISL_428917 | .................... | .................... | .................... | .................... | .................... | .................... |
| A | 9143 | hCoV-19/Russia/Novosibirsk-84101/2020\|EPI_ISL_4289 | .................... | .................... | .................... | .................... | .................... | .................... |
| A | 9144 | hCoV-19/Russia/Buryatia-87106/2020\|EPI_ISL_428919\| | .................... | .................... | .................... | .................... | .................... | .................... |
| A | 9145 | hCoV-19/Russia/Buryatia-87105/2020\|EPI_ISL_428920\| | .................... | .................... | .................... | .................... | .................... | .................... |
| A | 9146 | hCoV-19/Russia/Tver-72601/2020\|EPI_ISL_428921\|2020 | .................... | .................... | .................... | .................... | .................... | .................... |
| A | 9147 | hCoV-19/Russia/Sverdlovsk-86704/2020\|EPI_ISL_42892 | .................... | .................... | .................... | .................... | .................... | .................... |
| A | 9148 | hCoV-19/Russia/Sverdlovsk-84302/2020\|EPI_ISL_42892 | .................... | .................... | .................... | .................... | .................... | .................... |
| A | 9149 | hCoV-19/Russia/Chelyabinsk-85311/2020\|EPI_ISL_4289 | .................... | .................... | .................... | .................... | ....T............... | .................... |
| A | 9150 | hCoV-19/Poland/PL_P2/2020\|EPI_ISL_428925\|2020-03-2 | .................... | .................... | .................... | .................... | .................... | .................... |
| A | 9151 | hCoV-19/Poland/PL_P3/2020\|EPI_ISL_428926\|2020-03-2 | .................... | .................... | .................... | .................... | .................... | .................... |
| A | 9152 | hCoV-19/Poland/PL_P4/2020\|EPI_ISL_428927\|2020-03-2 | .................... | .................... | .................... | .................... | .................... | .................... |
| A | 9153 | hCoV-19/Poland/PL_P5/2020\|EPI_ISL_428928\|2020-03-3 | .................... | .................... | .................... | .................... | .................... | .................... |
| A | 9154 | hCoV-19/Poland/PL_P6/2020\|EPI_ISL_428929\|2020-03-2 | .................... | .................... | .................... | .................... | .................... | .................... |
| A | 9155 | hCoV-19/Poland/PL_P7/2020\|EPI_ISL_428930\|2020-03-2 | .................... | .................... | .................... | .................... | .................... | .................... |
| A | 9156 | hCoV-19/Poland/PL_P8/2020\|EPI_ISL_428931\|2020-03-2 | .................... | .................... | .................... | .................... | .................... | .................... |
| A | 9157 | hCoV-19/Poland/PL_P9/2020\|EPI_ISL_428932\|2020-03-2 | .................... | .................... | .................... | .................... | .................... | .................... |
| A | 9158 | hCoV-19/USA/WI-UW-259/2020\|EPI_ISL_428935\|2020-04- | .................... | .................... | .................... | .................... | .................... | .................... |
| A | 9159 | hCoV-19/USA/CA-CZB0104/2020\|EPI_ISL_428990\|2020-03 | .................... | .................... | .................... | .................... | .................... | .................... |

| | | | | | | | | |
|---|---|---|---|---|---|---|---|---|
| A | 9160 | hCoV-19/USA/CA-CZB071/2020\|EPI_ISL_428991\|2020-03- | ···················· | ···················· | ···················· | ···················· | ···················· | ···················· |
| A | 9161 | hCoV-19/USA/CA-CZB0138/2020\|EPI_ISL_428992\|2020-04 | ···················· | ···················· | ···················· | ···················· | ···················· | ···················· |
| A | 9162 | hCoV-19/USA/CA-CZB0157/2020\|EPI_ISL_428993\|2020-04 | ···················· | ···················· | ···················· | ···················· | ···················· | ···················· |
| A | 9163 | hCoV-19/USA/CA-CZB0703/2020\|EPI_ISL_428994\|2020-03 | ···················· | ···················· | ···················· | ···················· | ···················· | ···················· |
| A | 9164 | hCoV-19/USA/CA-CZB0133/2020\|EPI_ISL_428995\|2020-04 | ···················· | ···················· | ···················· | ···················· | ···················· | ···················· |
| A | 9165 | hCoV-19/USA/CA-CZB077/2020\|EPI_ISL_428996\|2020-03- | ···················· | ···················· | ···················· | ···················· | ···················· | ···················· |
| A | 9166 | hCoV-19/USA/CA-CZB015b/2020\|EPI_ISL_428997\|2020-03 | ···················· | ···················· | ···················· | ···················· | ···················· | ···················· |
| A | 9167 | hCoV-19/USA/CA-CZB084/2020\|EPI_ISL_428998\|2020-03- | ···················· | ···················· | ···················· | ···················· | ···················· | ···················· |
| A | 9168 | hCoV-19/USA/CA-CZB069/2020\|EPI_ISL_428999\|2020-03- | ···················· | ···················· | ···················· | ···················· | ···················· | ···················· |
| B | 1 | hCoV-19/USA/WA-S259/2020\|EPI_ISL_430113\|2020-03-04 | ···················· | ···················· | ···················· | ···················· | ···················· | ···················· |
| B | 2 | hCoV-19/USA/WA-S260/2020\|EPI_ISL_430114\|2020-03-12 | ···················· | ···················· | ···················· | ···················· | ···················· | ···················· |
| B | 3 | hCoV-19/USA/WA-S261/2020\|EPI_ISL_430115\|2020-03-30 | ···················· | ···················· | ···················· | ···················· | ···················· | ···················· |
| B | 4 | hCoV-19/USA/WA-S262/2020\|EPI_ISL_430116\|2020-03-16 | ···················· | ···················· | ···················· | ···················· | ···················· | ···················· |
| B | 5 | hCoV-19/USA/WA-S263/2020\|EPI_ISL_430117\|2020-03-05 | ···················· | ···················· | ···················· | ···················· | ···················· | ···················· |
| B | 6 | hCoV-19/USA/WA-S264/2020\|EPI_ISL_430118\|2020-03-03 | ···················· | ···················· | ···················· | ···················· | ···················· | ···················· |
| B | 7 | hCoV-19/USA/WA-S265/2020\|EPI_ISL_430119\|2020-03-05 | ···················· | ···················· | ···················· | ···················· | ···················· | ···················· |
| B | 8 | hCoV-19/USA/WA-S267/2020\|EPI_ISL_430121\|2020-03-31 | ···················· | ···················· | ···················· | ···················· | ···················· | ···················· |
| B | 9 | hCoV-19/USA/WA-S268/2020\|EPI_ISL_430122\|2020-03-05 | ···················· | ···················· | ···················· | ···················· | ···················· | ···················· |
| B | 10 | hCoV-19/USA/WA-S269/2020\|EPI_ISL_430123\|2020-03-10 | ···················· | ···················· | ···················· | ···················· | ···················· | ···················· |
| B | 11 | hCoV-19/USA/WA-S270/2020\|EPI_ISL_430124\|2020-03-06 | ···················· | ···················· | ···················· | ···················· | ···················· | ···················· |
| B | 12 | hCoV-19/USA/WA-S271/2020\|EPI_ISL_430125\|2020-03-31 | ···················· | ···················· | ···················· | ···················· | ···················· | ···················· |
| B | 13 | hCoV-19/USA/WA-S272/2020\|EPI_ISL_430126\|2020-02-26 | ···················· | ···················· | ···················· | ···················· | ···················· | ···················· |
| B | 14 | hCoV-19/USA/WA-S273/2020\|EPI_ISL_430127\|2020-03-12 | ···················· | ···················· | ···················· | ···················· | ···················· | ···················· |
| B | 15 | hCoV-19/USA/WA-S274/2020\|EPI_ISL_430128\|2020-03-08 | ···················· | ···················· | ···················· | ···················· | ···················· | ···················· |
| B | 16 | hCoV-19/USA/WA-S275/2020\|EPI_ISL_430129\|2020-03-06 | ···················· | ···················· | ···················· | ···················· | ···················· | ···················· |
| B | 17 | hCoV-19/USA/WA-S276/2020\|EPI_ISL_430130\|2020-04-01 | ···················· | ···················· | ···················· | ···················· | ···················· | ···················· |
| B | 18 | hCoV-19/USA/WA-S277/2020\|EPI_ISL_430131\|2020-02-22 | ···················· | ···················· | ···················· | ···················· | ···················· | ···················· |
| B | 19 | hCoV-19/USA/WA-S278/2020\|EPI_ISL_430132\|2020-03-17 | ···················· | ···················· | ···················· | ···················· | ···················· | ···················· |
| B | 20 | hCoV-19/USA/WA-S279/2020\|EPI_ISL_430133\|2020-03-04 | ···················· | ···················· | ···················· | ···················· | ···················· | ···················· |
| B | 21 | hCoV-19/USA/WA-S280/2020\|EPI_ISL_430134\|2020-03-09 | ···················· | ···················· | ···················· | ···················· | ···················· | ···················· |
| B | 22 | hCoV-19/USA/WA-S281/2020\|EPI_ISL_430135\|2020-02-29 | ···················· | ···················· | ···················· | ···················· | ···················· | ···················· |
| B | 23 | hCoV-19/USA/WA-S282/2020\|EPI_ISL_430136\|2020-03-10 | ···················· | ···················· | ···················· | ···················· | ···················· | ···················· |
| B | 24 | hCoV-19/USA/WA-S283/2020\|EPI_ISL_430137\|2020-03-21 | ···················· | ···················· | ···················· | ···················· | ···················· | ···················· |
| B | 25 | hCoV-19/USA/WA-S286/2020\|EPI_ISL_430140\|2020-03-15 | ···················· | ···················· | ···················· | ···················· | ···················· | ···················· |
| B | 26 | hCoV-19/USA/WA-S287/2020\|EPI_ISL_430141\|2020-03-06 | ···················· | ···················· | ···················· | ···················· | ···················· | ···················· |
| B | 27 | hCoV-19/USA/WA-S288/2020\|EPI_ISL_430142\|2020-03-06 | ···················· | ···················· | ···················· | ···················· | ···················· | ···················· |
| B | 28 | hCoV-19/USA/WA-S289/2020\|EPI_ISL_430143\|2020-03-23 | ···················· | ···················· | ···················· | ···················· | ···················· | ···················· |
| B | 29 | hCoV-19/USA/WA-S292/2020\|EPI_ISL_430146\|2020-03-05 | ···················· | ···················· | ···················· | ···················· | ···················· | ···················· |
| B | 30 | hCoV-19/USA/WA-S293/2020\|EPI_ISL_430147\|2020-04-01 | ···················· | ···················· | ···················· | ···················· | ···················· | ···················· |
| B | 31 | hCoV-19/USA/WA-S294/2020\|EPI_ISL_430148\|2020-03-23 | ···················· | ···················· | ···················· | ···················· | ···················· | ···················· |
| B | 32 | hCoV-19/USA/WA-S295/2020\|EPI_ISL_430149\|2020-03-31 | ···················· | ···················· | ···················· | ···················· | ···················· | ···················· |
| B | 33 | hCoV-19/USA/WA-S296/2020\|EPI_ISL_430150\|2020-03-13 | ···················· | ···················· | ···················· | A··················· | ···················· | ···················· |
| B | 34 | hCoV-19/USA/WA-S297/2020\|EPI_ISL_430151\|2020-03-13 | ···················· | ···················· | ···················· | ···················· | ···················· | ···················· |
| B | 35 | hCoV-19/USA/WA-S298/2020\|EPI_ISL_430152\|2020-03-05 | ···················· | ···················· | ···················· | ···················· | ···················· | ···················· |
| B | 36 | hCoV-19/USA/WA-S299/2020\|EPI_ISL_430153\|2020-03-31 | ···················· | ···················· | ···················· | ···················· | ···················· | ···················· |
| B | 37 | hCoV-19/USA/WA-S300/2020\|EPI_ISL_430154\|2020-03-25 | ···················· | ···················· | ···················· | ···················· | ···················· | ···················· |
| B | 38 | hCoV-19/USA/WA-S301/2020\|EPI_ISL_430155\|2020-03-14 | ···················· | ···················· | ···················· | ···················· | ···················· | ···················· |
| B | 40 | hCoV-19/USA/WA-S123/2020\|EPI_ISL_430160\|2020-02-29 | ···················· | ···················· | ···················· | ···················· | ···················· | ···················· |
| B | 41 | hCoV-19/USA/WA-S124/2020\|EPI_ISL_430161\|2020-03-13 | ···················· | ···················· | ···················· | ···················· | ···················· | ···················· |
| B | 42 | hCoV-19/USA/WA-S125/2020\|EPI_ISL_430162\|2020-03-13 | ···················· | ···················· | ···················· | ···················· | ···················· | ···················· |
| B | 43 | hCoV-19/USA/WA-S126/2020\|EPI_ISL_430163\|2020-03-13 | ···················· | ···················· | ···················· | ···················· | ···················· | ···················· |
| B | 44 | hCoV-19/USA/WA-S127/2020\|EPI_ISL_430164\|2020-03-13 | ···················· | ···················· | ···················· | ···················· | ···················· | ···················· |
| B | 45 | hCoV-19/USA/WA-S128/2020\|EPI_ISL_430165\|2020-03-13 | ···················· | ···················· | ···················· | ···················· | ···················· | ···················· |
| B | 46 | hCoV-19/USA/WA-S129/2020\|EPI_ISL_430166\|2020-03-13 | ···················· | ···················· | ···················· | ···················· | ···················· | ···················· |
| B | 47 | hCoV-19/USA/WA-S130/2020\|EPI_ISL_430167\|2020-03-13 | ···················· | ···················· | ···················· | ···················· | ···················· | ···················· |
| B | 48 | hCoV-19/USA/WA-S131/2020\|EPI_ISL_430168\|2020-03-13 | ···················· | ···················· | ···················· | ···················· | ···················· | ···················· |
| B | 49 | hCoV-19/USA/WA-S132/2020\|EPI_ISL_430169\|2020-03-13 | ···················· | ···················· | ···················· | ···················· | ···················· | ···················· |
| B | 50 | hCoV-19/USA/WA-S133/2020\|EPI_ISL_430170\|2020-03-13 | ···················· | ···················· | ···················· | ···················· | ···················· | ···················· |
| B | 51 | hCoV-19/USA/WA-S134/2020\|EPI_ISL_430171\|2020-03-13 | ···················· | ···················· | ···················· | ···················· | ···················· | ···················· |
| B | 52 | hCoV-19/USA/WA-S135/2020\|EPI_ISL_430172\|2020-03-13 | ···················· | ···················· | ···················· | ···················· | ···················· | ···················· |
| B | 53 | hCoV-19/USA/WA-S136/2020\|EPI_ISL_430173\|2020-03-13 | ···················· | ···················· | ···················· | ···················· | ···················· | ···················· |
| B | 54 | hCoV-19/USA/WA-S137/2020\|EPI_ISL_430174\|2020-03-13 | ···················· | ···················· | ···················· | ···················· | ···················· | ···················· |
| B | 55 | hCoV-19/USA/WA-S138/2020\|EPI_ISL_430175\|2020-03-12 | ···················· | ···················· | ···················· | ···················· | ···················· | ···················· |
| B | 56 | hCoV-19/USA/WA-S139/2020\|EPI_ISL_430176\|2020-03-12 | ···················· | ···················· | ···················· | ···················· | ···················· | ···················· |
| B | 57 | hCoV-19/USA/WA-S140/2020\|EPI_ISL_430177\|2020-03-12 | ···················· | ···················· | ···················· | ···················· | ···················· | ···················· |
| B | 58 | hCoV-19/USA/WA-S141/2020\|EPI_ISL_430178\|2020-03-13 | ···················· | ···················· | ···················· | ···················· | ···················· | ···················· |
| B | 59 | hCoV-19/England/NOTT-10FC84/2020\|EPI_ISL_432706\|20 | ···················· | ···················· | ···················· | ···················· | ···················· | ···················· |
| B | 60 | hCoV-19/England/SHEF-C18E5/2020\|EPI_ISL_432732\|202 | ···················· | ···················· | ···················· | ···················· | ···················· | ···················· |
| B | 61 | hCoV-19/England/SHEF-C1B64/2020\|EPI_ISL_432734\|202 | ···················· | ···················· | ···················· | ···················· | ···················· | ···················· |
| B | 62 | hCoV-19/England/SHEF-C2222/2020\|EPI_ISL_432742\|202 | ···················· | ···················· | ···················· | ···················· | ···················· | ···················· |
| B | 63 | hCoV-19/England/SHEF-C2310/2020\|EPI_ISL_432744\|202 | ···················· | ···················· | ···················· | ···················· | ···················· | ···················· |
| B | 64 | hCoV-19/England/SHEF-C2626/2020\|EPI_ISL_432746\|202 | ···················· | ···················· | ···················· | ···················· | ···················· | ···················· |
| B | 65 | hCoV-19/England/SHEF-C2CF7/2020\|EPI_ISL_432753\|202 | ···················· | ···················· | ···················· | ···················· | ···················· | ···················· |
| B | 66 | hCoV-19/England/SHEF-C3115/2020\|EPI_ISL_432762\|202 | ···················· | ···················· | ···················· | ···················· | ···················· | ···················· |
| B | 67 | hCoV-19/England/SHEF-C3212/2020\|EPI_ISL_432766\|202 | ···················· | ···················· | ···················· | ···················· | ···················· | ···················· |
| B | 68 | hCoV-19/England/SHEF-C32C7/2020\|EPI_ISL_432768\|202 | ···················· | ···················· | ···················· | ···················· | ···················· | ···················· |
| B | 69 | hCoV-19/England/SHEF-C384D/2020\|EPI_ISL_432777\|202 | ···················· | ···················· | ···················· | ···················· | ···················· | ···················· |
| B | 70 | hCoV-19/England/SHEF-C3CC9/2020\|EPI_ISL_432780\|202 | ···················· | ···················· | ···················· | ···················· | ···················· | ···················· |

```
B   71 hCoV-19/England/SHEF-C426C/2020|EPI_ISL_432786|202   ····················  ····················  ····················  ····················  ····················  ····················
B   72 hCoV-19/England/SHEF-C4493/2020|EPI_ISL_432788|202   ····················  ····················  ····················  ····················  ····················  ····················
B   73 hCoV-19/England/SHEF-C45BE/2020|EPI_ISL_432790|202   ····················  ····················  ····················  ····················  ····················  ····················
B   74 hCoV-19/England/SHEF-C51E6/2020|EPI_ISL_432799|202   ····················  ····················  ····················  ····················  ····················  ····················
B   75 hCoV-19/Spain/VC-FISABIO-187/2020|EPI_ISL_436282|2   ····················  ····················  ····················  ····················  ····················  ····················
B   76 hCoV-19/Spain/VC-FISABIO-188/2020|EPI_ISL_436283|2   ····················  ····················  ····················  ····················  ····················  ····················
B   77 hCoV-19/Spain/VC-FISABIO-189/2020|EPI_ISL_436284|2   ····················  ····················  ····················  ····················  ····················  ····················
B   78 hCoV-19/Spain/VC-FISABIO-190/2020|EPI_ISL_436285|2   ····················  ····················  ····················  ····················  ····················  ····················
B   79 hCoV-19/Spain/VC-FISABIO-192/2020|EPI_ISL_436287|2   ····················  ····················  ····················  ····················  ····················  ····················
B   80 hCoV-19/Spain/VC-FISABIO-193/2020|EPI_ISL_436288|2   ····················  ····················  ····················  ····················  ····················  ····················
B   81 hCoV-19/Spain/VC-FISABIO-195/2020|EPI_ISL_436290|2   ····················  ····················  ····················  ····················  ····················  ····················
B   82 hCoV-19/Spain/VC-FISABIO-196/2020|EPI_ISL_436291|2   ····················  ····················  ····················  ····················  ····················  ····················
B   83 hCoV-19/Spain/VC-FISABIO-197/2020|EPI_ISL_436292|2   ····················  ····················  ····················  ····················  ····················  ····················
B   84 hCoV-19/Spain/VC-FISABIO-200/2020|EPI_ISL_436295|2   ····················  ····················  ····················  ····················  ····················  ····················
B   86 hCoV-19/England/CAMB-7BF30/2020|EPI_ISL_442101|202   ····················  ····················  ····················  ····················  ····················  ····················
B   87 hCoV-19/England/CAMB-7C89E/2020|EPI_ISL_442102|202   ····················  ····················  ····················  ····················  ····················  ····················
B   88 hCoV-19/England/CAMB-7C782/2020|EPI_ISL_442103|202   ····················  ····················  ····················  ····················  ····················  ····················
B   89 hCoV-19/England/CAMB-7BDBE/2020|EPI_ISL_442104|202   ····················  ····················  ····················  ····················  ····················  ····················
B   90 hCoV-19/England/CAMB-7BBC3/2020|EPI_ISL_442105|202   ····················  ····················  ····················  ····················  ····················  ····················
B   91 hCoV-19/England/CAMB-7D666/2020|EPI_ISL_442106|202   ····················  ····················  ····················  ····················  ····················  ····················
B   92 hCoV-19/England/CAMB-7BD18/2020|EPI_ISL_442107|202   ····················  ····················  ····················  ····················  ····················  ····················
B   93 hCoV-19/England/CAMB-7D138/2020|EPI_ISL_442108|202   ····················  ····················  ····················  ····················  ····················  ····················
B   94 hCoV-19/England/CAMB-7BE42/2020|EPI_ISL_442109|202   ····················  ····················  ····················  ····················  ····················  ····················
B   95 hCoV-19/England/CAMB-7D03B/2020|EPI_ISL_442111|202   ····················  ····················  ····················  ····················  ····················  ····················
B   96 hCoV-19/England/CAMB-7BBE1/2020|EPI_ISL_442112|202   ····················  ····················  ····················  ····················  ····················  ····················
B   97 hCoV-19/England/CAMB-7D5C3/2020|EPI_ISL_442113|202   ····················  ····················  ····················  ····················  ····················  ····················
B   98 hCoV-19/England/CAMB-7C9B9/2020|EPI_ISL_442114|202   ····················  ····················  ····················  ····················  ····················  ····················
B   99 hCoV-19/England/CAMB-7C9F5/2020|EPI_ISL_442116|202   ····················  ····················  ····················  ····················  ····················  ····················
B  100 hCoV-19/England/CAMB-7B8DB/2020|EPI_ISL_442117|202   ····················  ····················  ····················  ····················  ····················  ····················
B  101 hCoV-19/England/CAMB-7C96E/2020|EPI_ISL_442120|202   ····················  ····················  ····················  ····················  ····················  ····················
B  102 hCoV-19/England/CAMB-7CB77/2020|EPI_ISL_442121|202   ····················  ····················  ····················  ····················  ····················  ····················
B  103 hCoV-19/England/CAMB-7BD54/2020|EPI_ISL_442122|202   ····················  ····················  ····················  ····················  ····················  ····················
B  104 hCoV-19/England/CAMB-7D262/2020|EPI_ISL_442123|202   ····················  ····················  ····················  ····················  ····················  ····················
B  105 hCoV-19/England/CAMB-7C8DA/2020|EPI_ISL_442124|202   ····················  ····················  ····················  ····················  ····················  ····················
B  106 hCoV-19/England/CAMB-7E094/2020|EPI_ISL_442125|202   ····················  ·A··················  ····················  ····················  ····················  ····················
B  107 hCoV-19/England/CAMB-7BE33/2020|EPI_ISL_442126|202   ····················  ····················  ····················  ····················  ····················  ····················
B  108 hCoV-19/England/CAMB-7D60C/2020|EPI_ISL_442129|202   ····················  ····················  ····················  ····················  ····················  ····················
B  109 hCoV-19/England/CAMB-7BB5A/2020|EPI_ISL_442130|202   ····················  ····················  ····················  ····················  ····················  ····················
B  110 hCoV-19/England/CAMB-7E0A3/2020|EPI_ISL_442133|202   ····················  ····················  ····················  ····················  ····················  ····················
B  111 hCoV-19/England/CAMB-7BDEB/2020|EPI_ISL_442134|202   ····················  ····················  ····················  ····················  ····················  ····················
B  112 hCoV-19/England/CAMB-7C9D7/2020|EPI_ISL_442136|202   ····················  ····················  ····················  ····················  ····················  ····················
B  113 hCoV-19/England/CAMB-7BCDF/2020|EPI_ISL_442139|202   ····················  ····················  ····················  ····················  ····················  ····················
B  114 hCoV-19/England/CAMB-7CAD4/2020|EPI_ISL_442140|202   ····················  ····················  ····················  ····················  ····················  ····················
B  115 hCoV-19/England/CAMB-7D174/2020|EPI_ISL_442142|202   ····················  ····················  ····················  ····················  ····················  ····················
B  116 hCoV-19/England/CAMB-7CBFF/2020|EPI_ISL_442143|202   ····················  ····················  ····················  ····················  ····················  ····················
B  117 hCoV-19/England/CAMB-7BCFD/2020|EPI_ISL_442144|202   ····················  ····················  ····················  ····················  ····················  ····················
B  118 hCoV-19/England/CAMB-7BC84/2020|EPI_ISL_442146|202   ····················  ····················  ····················  ····················  ····················  ····················
B  119 hCoV-19/England/CAMB-7CA2F/2020|EPI_ISL_442147|202   ····················  ····················  ····················  ····················  ····················  ····················
B  120 hCoV-19/England/CAMB-7D244/2020|EPI_ISL_442149|202   ····················  ····················  ····················  ····················  ····················  ····················
B  121 hCoV-19/England/CAMB-7CC74/2020|EPI_ISL_442150|202   ····················  ····················  ····················  ····················  ····················  ····················
B  122 hCoV-19/England/CAMB-7BED9/2020|EPI_ISL_442154|202   ····················  ····················  ····················  ····················  ····················  ····················
B  123 hCoV-19/England/CAMB-7CC92/2020|EPI_ISL_442157|202   ····················  ····················  ····················  ····················  ····················  ····················
B  124 hCoV-19/England/CAMB-830AB/2020|EPI_ISL_443402|202   ····················  ····················  ····················  ····················  ····················  ····················
B  125 hCoV-19/England/CAMB-843DE/2020|EPI_ISL_443403|202   ····················  ····················  ····················  ····················  ····················  ····················
B  126 hCoV-19/England/CAMB-827F2/2020|EPI_ISL_443404|202   ····················  ····················  ····················  ····················  ····················  ····················
B  127 hCoV-19/England/CAMB-8459C/2020|EPI_ISL_443406|202   ····················  ····················  ····················  ····················  ····················  ····················
B  128 hCoV-19/England/CAMB-1AA2D4/2020|EPI_ISL_443408|20   ····················  ····················  ····················  ····················  ····················  ····················
B  129 hCoV-19/England/CAMB-838B2/2020|EPI_ISL_443409|202   ····················  ····················  ····················  ····················  ····················  ····················
B  130 hCoV-19/England/CAMB-1AA3FF/2020|EPI_ISL_443410|20   ····················  ····················  ····················  ····················  ····················  ····················
B  131 hCoV-19/England/CAMB-1AA395/2020|EPI_ISL_443411|20   ····················  ····················  ····················  ····················  ····················  ····················
B  132 hCoV-19/England/CAMB-1AA2C5/2020|EPI_ISL_443414|20   ····················  ····················  ····················  ····················  ····················  ····················
B  133 hCoV-19/England/CAMB-833DF/2020|EPI_ISL_443415|202   ····················  ····················  ····················  ····················  ····················  ····················
B  134 hCoV-19/England/CAMB-833A2/2020|EPI_ISL_443416|202   ····················  ····················  ····················  ····················  ····················  ····················
B  135 hCoV-19/England/CAMB-828D1/2020|EPI_ISL_443417|202   ····················  ····················  ····················  ····················  ····················  ····················
B  136 hCoV-19/England/CAMB-845D8/2020|EPI_ISL_443419|202   ····················  ····················  ····················  ····················  ····················  ····················
B  137 hCoV-19/England/CAMB-1AA3D1/2020|EPI_ISL_443421|20   ····················  ····················  ····················  ····················  ····················  ····················
B  138 hCoV-19/England/CAMB-82F72/2020|EPI_ISL_443425|202   ····················  ····················  ····················  ····················  ····················  ····················
B  139 hCoV-19/England/CAMB-82552/2020|EPI_ISL_443426|202   ····················  ····················  ····················  ····················  ····················  ····················
B  140 hCoV-19/England/CAMB-82D3C/2020|EPI_ISL_443427|202   ····················  ····················  ····················  ····················  ····················  ····················
B  141 hCoV-19/England/CAMB-8225B/2020|EPI_ISL_443428|202   ····················  ····················  ····················  ····················  ····················  ····················
B  142 hCoV-19/England/CAMB-831E4/2020|EPI_ISL_443429|202   ····················  ····················  ····················  ····················  ····················  ····················
B  143 hCoV-19/England/CAMB-84444/2020|EPI_ISL_443434|202   ····················  ····················  ····················  ····················  ····················  ····················
B  144 hCoV-19/England/CAMB-83788/2020|EPI_ISL_443436|202   ····················  ····················  ····················  ····················  ····················  ····················
B  145 hCoV-19/England/CAMB-83005/2020|EPI_ISL_443437|202   ····················  ····················  ····················  ····················  ····················  ····················
B  146 hCoV-19/England/CAMB-83366/2020|EPI_ISL_443440|202   ····················  ····················  ····················  ····················  ····················  ····················
B  147 hCoV-19/England/CAMB-1AA4ED/2020|EPI_ISL_443441|20   ····················  ····················  ····················  ····················  ····················  ····················
B  148 hCoV-19/England/CAMB-83560/2020|EPI_ISL_443444|202   ····················  ····················  ····················  ····················  ····················  ····················
B  149 hCoV-19/England/CAMB-843CF/2020|EPI_ISL_443445|202   ····················  ····················  ····················  ····················  ····················  ····················
```

| | | | | | | | |
|---|---|---|---|---|---|---|---|
| B | 150 hCoV-19/England/CAMB-83797/2020\|EPI_ISL_443446\|202 | ···················· | ···················· | ···················· | ···················· | ···················· | ···················· |
| B | 151 hCoV-19/England/CAMB-834DC/2020\|EPI_ISL_443447\|202 | ···················· | ···················· | ···················· | ···················· | ···················· | ···················· |
| B | 152 hCoV-19/England/CAMB-1AA3E0/2020\|EPI_ISL_443448\|20 | ···················· | ···················· | ···················· | ···················· | ···················· | ···················· |
| B | 153 hCoV-19/England/CAMB-82482/2020\|EPI_ISL_443450\|202 | ···················· | ···················· | ···················· | ···················· | ···················· | ···················· |
| B | 154 hCoV-19/England/CAMB-8224C/2020\|EPI_ISL_443452\|202 | ···················· | ······R············· | ···················· | ···················· | ···················· | ···················· |
| B | 155 hCoV-19/England/CAMB-83876/2020\|EPI_ISL_443453\|202 | ···················· | ···················· | ···················· | ···················· | ···················· | ···················· |
| B | 156 hCoV-19/England/CAMB-8422C/2020\|EPI_ISL_443454\|202 | ···················· | ···················· | ···················· | ···················· | ···················· | ···················· |
| B | 157 hCoV-19/England/CAMB-83515/2020\|EPI_ISL_443455\|202 | ···················· | ···················· | ···················· | ···················· | ···················· | ···················· |
| B | 158 hCoV-19/England/CAMB-826C8/2020\|EPI_ISL_443456\|202 | ···················· | ···················· | ···················· | ···················· | ···················· | ···················· |
| B | 159 hCoV-19/England/CAMB-1AA562/2020\|EPI_ISL_443457\|20 | ···················· | ···················· | ···················· | ···················· | ···················· | ···················· |
| B | 160 hCoV-19/England/CAMB-82491/2020\|EPI_ISL_443458\|202 | ···················· | ···················· | ···················· | ···················· | ···················· | ···················· |
| B | 161 hCoV-19/England/CAMB-82507/2020\|EPI_ISL_443459\|202 | ···················· | ···················· | ···················· | ···················· | ···················· | ···················· |
| B | 162 hCoV-19/England/CAMB-842D1/2020\|EPI_ISL_443460\|202 | ···················· | ···················· | ···················· | ···················· | ···················· | ···················· |
| B | 163 hCoV-19/England/CAMB-8233A/2020\|EPI_ISL_443461\|202 | ···················· | ···················· | ···················· | ···················· | ···················· | ···················· |
| B | 164 hCoV-19/England/CAMB-8358E/2020\|EPI_ISL_443463\|202 | ···················· | ···················· | ···················· | ···················· | ···················· | ···················· |
| B | 165 hCoV-19/England/CAMB-827A7/2020\|EPI_ISL_443464\|202 | ···················· | ···················· | ···················· | ···················· | ···················· | ···················· |
| B | 166 hCoV-19/England/CAMB-823EF/2020\|EPI_ISL_443465\|202 | ···················· | ···················· | ···················· | ···················· | ···················· | ···················· |
| B | 167 hCoV-19/England/CAMB-1AA474/2020\|EPI_ISL_443466\|20 | ···················· | ·A·················· | ···················· | ···················· | ···················· | ···················· |
| B | 168 hCoV-19/England/CAMB-82D1E/2020\|EPI_ISL_443467\|202 | ···················· | ···················· | ···················· | ···················· | ···················· | ···················· |
| B | 169 hCoV-19/England/CAMB-84462/2020\|EPI_ISL_443469\|202 | ···················· | ···················· | ···················· | ···················· | ···················· | ···················· |
| B | 170 hCoV-19/England/CAMB-837D3/2020\|EPI_ISL_443470\|202 | ···················· | ···················· | ···················· | ···················· | ···················· | ···················· |
| B | 171 hCoV-19/England/CAMB-824EC/2020\|EPI_ISL_443472\|202 | ···················· | ···················· | ···················· | ···················· | ···················· | ···················· |
| B | 172 hCoV-19/England/CAMB-833FD/2020\|EPI_ISL_443473\|202 | ···················· | ···················· | ···················· | ···················· | ···················· | ···················· |
| B | 173 hCoV-19/England/CAMB-8374C/2020\|EPI_ISL_443474\|202 | ···················· | ···················· | ···················· | ···················· | ···················· | ···················· |
| B | 174 hCoV-19/England/CAMB-842A4/2020\|EPI_ISL_443476\|202 | ···················· | ···················· | ···················· | ···················· | ···················· | ···················· |
| B | 175 hCoV-19/England/CAMB-8376A/2020\|EPI_ISL_443477\|202 | ···················· | ···················· | ···················· | ···················· | ···················· | ···················· |
| B | 176 hCoV-19/England/CAMB-833B1/2020\|EPI_ISL_443478\|202 | ···················· | ···················· | ···················· | ···················· | ···················· | ···················· |
| B | 177 hCoV-19/England/CAMB-822E2/2020\|EPI_ISL_443481\|202 | ···················· | ···················· | ···················· | ···················· | ···················· | ···················· |
| B | 178 hCoV-19/England/CAMB-82FDC/2020\|EPI_ISL_443482\|202 | ···················· | ···················· | ···················· | ···················· | ···················· | ···················· |
| B | 179 hCoV-19/England/CAMB-82D0F/2020\|EPI_ISL_443483\|202 | ···················· | ···················· | ···················· | ···················· | ···················· | ···················· |
| B | 180 hCoV-19/England/CAMB-8223D/2020\|EPI_ISL_443487\|202 | ···················· | ···················· | ···················· | ···················· | ···················· | ···················· |
| B | 181 hCoV-19/England/CAMB-83472/2020\|EPI_ISL_443488\|202 | ···················· | ···················· | ···················· | ···················· | ···················· | ···················· |
| B | 182 hCoV-19/USA/NY-NYUMC593/2020\|EPI_ISL_444710\|2020-0 | ···················· | ···················· | ···················· | ···················· | ···················· | ···················· |
| B | 183 hCoV-19/USA/NY-NYUMC594/2020\|EPI_ISL_444711\|2020-0 | ···················· | ···················· | ···················· | ···················· | ···················· | ···················· |
| B | 184 hCoV-19/USA/NY-NYUMC595/2020\|EPI_ISL_444712\|2020-0 | ···················· | ···················· | ···················· | ···················· | ···················· | ···················· |
| B | 185 hCoV-19/USA/NY-NYUMC596/2020\|EPI_ISL_444713\|2020-0 | ···················· | ···················· | ···················· | ···················· | ···················· | ···················· |
| B | 186 hCoV-19/USA/NY-NYUMC597/2020\|EPI_ISL_444714\|2020-0 | ···················· | ···················· | ···················· | ···················· | ···················· | ···················· |
| B | 187 hCoV-19/USA/NY-NYUMC598/2020\|EPI_ISL_444715\|2020-0 | ···················· | ···················· | ···················· | ···················· | ···················· | ···················· |
| B | 188 hCoV-19/USA/NY-NYUMC599/2020\|EPI_ISL_444716\|2020-0 | ···················· | ···················· | ···················· | ···················· | ···················· | ···················· |
| B | 189 hCoV-19/USA/NY-NYUMC600/2020\|EPI_ISL_444717\|2020-0 | ···················· | ···················· | ···················· | ···················· | ···················· | ···················· |
| B | 190 hCoV-19/USA/NY-NYUMC601/2020\|EPI_ISL_444718\|2020-0 | ···················· | ···················· | ···················· | ···················· | ···················· | ···················· |
| B | 191 hCoV-19/USA/NY-NYUMC602/2020\|EPI_ISL_444719\|2020-0 | ···················· | ···················· | ···················· | ···················· | ···················· | ···················· |
| B | 192 hCoV-19/USA/NY-NYUMC603/2020\|EPI_ISL_444720\|2020-0 | ···················· | ···················· | ···················· | ···················· | ···················· | ···················· |
| B | 193 hCoV-19/USA/NY-NYUMC605/2020\|EPI_ISL_444722\|2020-0 | ···················· | ···················· | ···················· | ···················· | ···················· | ···················· |
| B | 194 hCoV-19/USA/NY-NYUMC606/2020\|EPI_ISL_444723\|2020-0 | ···················· | ···················· | ···················· | ···················· | ···················· | ···················· |
| B | 195 hCoV-19/USA/NY-NYUMC607/2020\|EPI_ISL_444724\|2020-0 | ···················· | ···················· | ···················· | ···················· | ···················· | ···················· |
| B | 196 hCoV-19/USA/NY-NYUMC612/2020\|EPI_ISL_444729\|2020-0 | ···················· | ···················· | ···················· | ···················· | ···················· | ···················· |
| B | 197 hCoV-19/USA/NY-NYUMC613/2020\|EPI_ISL_444730\|2020-0 | ···················· | ···················· | ···················· | ···················· | ···················· | ···················· |
| B | 198 hCoV-19/USA/NY-NYUMC614/2020\|EPI_ISL_444731\|2020-0 | ···················· | ···················· | ···················· | ···················· | ···················· | ···················· |
| B | 199 hCoV-19/USA/NY-NYUMC615/2020\|EPI_ISL_444732\|2020-0 | ···················· | ···················· | ···················· | ···················· | ···················· | ···················· |
| B | 200 hCoV-19/USA/NY-NYUMC616/2020\|EPI_ISL_444733\|2020-0 | ···················· | ···················· | ···················· | ···················· | ···················· | ···················· |
| B | 201 hCoV-19/USA/NY-NYUMC617/2020\|EPI_ISL_444734\|2020-0 | ···················· | ···················· | ···················· | ···················· | ···················· | ···················· |
| B | 202 hCoV-19/USA/NY-NYUMC618/2020\|EPI_ISL_444735\|2020-0 | ···················· | ···················· | ···················· | ···················· | ···················· | ···················· |
| B | 203 hCoV-19/USA/NY-NYUMC619/2020\|EPI_ISL_444736\|2020-0 | ···················· | ···················· | ···················· | ···················· | ···················· | ···················· |
| B | 204 hCoV-19/USA/NY-NYUMC620/2020\|EPI_ISL_444737\|2020-0 | ···················· | ····A··············· | ···················· | ···················· | ···················· | ···················· |
| B | 205 hCoV-19/USA/NY-NYUMC621/2020\|EPI_ISL_444738\|2020-0 | ···················· | ···················· | ···················· | ···················· | ···················· | ···················· |
| B | 206 hCoV-19/USA/NY-NYUMC622/2020\|EPI_ISL_444739\|2020-0 | ···················· | ···················· | ···················· | ···················· | ···················· | ···················· |
| B | 207 hCoV-19/USA/NY-NYUMC623/2020\|EPI_ISL_444740\|2020-0 | ···················· | ···················· | ···················· | ···················· | ···················· | ···················· |
| B | 208 hCoV-19/USA/NY-NYUMC624/2020\|EPI_ISL_444741\|2020-0 | ···················· | ···················· | ···················· | ···················· | ···················· | ···················· |
| B | 209 hCoV-19/USA/NJ-NYUMC625/2020\|EPI_ISL_444742\|2020-0 | ···················· | ···················· | ···················· | ···················· | ···················· | ···················· |
| B | 210 hCoV-19/USA/NY-NYUMC626/2020\|EPI_ISL_444743\|2020-0 | ···················· | ···················· | ···················· | ···················· | ···················· | ···················· |
| B | 211 hCoV-19/USA/NJ-NYUMC627/2020\|EPI_ISL_444744\|2020-0 | ···················· | ···················· | ···················· | ···················· | ···················· | ···················· |
| B | 212 hCoV-19/USA/NY-NYUMC628/2020\|EPI_ISL_444745\|2020-0 | ···················· | ···················· | ···················· | ···················· | ···················· | ···················· |
| B | 213 hCoV-19/USA/NY-NYUMC629/2020\|EPI_ISL_444746\|2020-0 | ···················· | ···················· | ···················· | ···················· | ···················· | ···················· |
| B | 214 hCoV-19/USA/NY-NYUMC630/2020\|EPI_ISL_444747\|2020-0 | ···················· | ···················· | ···················· | ···················· | ···················· | ···················· |
| B | 215 hCoV-19/USA/NY-NYUMC631/2020\|EPI_ISL_444748\|2020-0 | ···················· | ···················· | ···················· | ···················· | ···················· | ···················· |
| B | 216 hCoV-19/USA/NY-NYUMC632/2020\|EPI_ISL_444749\|2020-0 | ···················· | ···················· | ···················· | ···················· | ···················· | ···················· |
| B | 217 hCoV-19/USA/NY-NYUMC633/2020\|EPI_ISL_444750\|2020-0 | ···················· | ···················· | ···················· | ···················· | ···················· | ···················· |
| B | 218 hCoV-19/USA/NY-NYUMC634/2020\|EPI_ISL_444751\|2020-0 | ···················· | ···················· | ···················· | ···················· | ···················· | ···················· |
| B | 219 hCoV-19/USA/NY-NYUMC635/2020\|EPI_ISL_444752\|2020-0 | ···················· | ···················· | ···················· | ···················· | ···················· | ···················· |
| B | 220 hCoV-19/USA/NY-NYUMC636/2020\|EPI_ISL_444753\|2020-0 | ···················· | ···················· | ···················· | ···················· | ···················· | ···················· |
| B | 221 hCoV-19/USA/NY-NYUMC637/2020\|EPI_ISL_444754\|2020-0 | ···················· | ···················· | ···················· | ···················· | ···················· | ···················· |
| B | 222 hCoV-19/USA/NJ-NYUMC638/2020\|EPI_ISL_444755\|2020-0 | ···················· | ···················· | ···················· | ···················· | ···················· | ···················· |
| B | 223 hCoV-19/USA/NY-NYUMC639/2020\|EPI_ISL_444756\|2020-0 | ···················· | ···················· | ···················· | ···················· | ···················· | ···················· |
| B | 224 hCoV-19/USA/NY-NYUMC640/2020\|EPI_ISL_444757\|2020-0 | ···················· | ···················· | ···················· | ···················· | ···················· | ···················· |
| B | 225 hCoV-19/USA/NY-NYUMC641/2020\|EPI_ISL_444758\|2020-0 | ···················· | ···················· | ···················· | ···················· | ···················· | ···················· |
| B | 226 hCoV-19/USA/NY-NYUMC642/2020\|EPI_ISL_444759\|2020-0 | ···················· | ···················· | ···················· | ···················· | ···················· | ···················· |
| B | 227 hCoV-19/USA/NY-NYUMC643/2020\|EPI_ISL_444760\|2020-0 | ···················· | ···················· | ···················· | ···················· | ···················· | ···················· |

| | | | | | | | | |
|---|---|---|---|---|---|---|---|---|
| B | 228 | hCoV-19/USA/NY-NYUMC644/2020\|EPI_ISL_444761\|2020-0 | .................... | .................... | ..................... | .................... | .................... | .................... |
| B | 229 | hCoV-19/USA/NY-NYUMC645/2020\|EPI_ISL_444762\|2020-0 | .................... | .................... | ..................... | .................... | .................... | .................... |
| B | 230 | hCoV-19/USA/NY-NYUMC646/2020\|EPI_ISL_444763\|2020-0 | .................... | .................... | ..................... | .................... | .................... | .................... |
| B | 231 | hCoV-19/USA/NY-NYUMC647/2020\|EPI_ISL_444764\|2020-0 | .................... | .................... | ..................... | .................... | .................... | .................... |
| B | 232 | hCoV-19/USA/NY-NYUMC648/2020\|EPI_ISL_444765\|2020-0 | .................... | .................... | ..................... | .................... | .................... | .................... |
| B | 233 | hCoV-19/USA/NY-NYUMC649/2020\|EPI_ISL_444766\|2020-0 | .................... | .................... | ..................... | .................... | .................... | .................... |
| B | 234 | hCoV-19/USA/NY-NYUMC650/2020\|EPI_ISL_444767\|2020-0 | .................... | .................... | ..................... | .................... | .................... | .................... |
| B | 235 | hCoV-19/USA/NY-NYUMC651/2020\|EPI_ISL_444768\|2020-0 | .................... | .................... | ..................... | .................... | .................... | .................... |
| B | 236 | hCoV-19/USA/NY-NYUMC652/2020\|EPI_ISL_444769\|2020-0 | .................... | .................... | ..................... | .................... | .................... | .................... |
| B | 237 | hCoV-19/USA/NY-NYUMC653/2020\|EPI_ISL_444770\|2020-0 | .................... | .................... | ..................... | .................... | .................... | .................... |
| B | 238 | hCoV-19/USA/NY-NYUMC654/2020\|EPI_ISL_444771\|2020-0 | .................... | .................... | ..................... | .................... | .................... | .................... |
| B | 239 | hCoV-19/USA/NY-NYUMC655/2020\|EPI_ISL_444772\|2020-0 | .................... | .................... | ..................... | .................... | .................... | .................... |
| B | 240 | hCoV-19/USA/NY-NYUMC656/2020\|EPI_ISL_444773\|2020-0 | .................... | .................... | ..................... | .................... | .................... | .................... |
| B | 241 | hCoV-19/USA/NY-NYUMC657/2020\|EPI_ISL_444774\|2020-0 | .................... | .................... | ..................... | .................... | .................... | .................... |
| B | 242 | hCoV-19/USA/NY-NYUMC658/2020\|EPI_ISL_444775\|2020-0 | .................... | .................... | ..................... | .................... | .................... | .................... |
| B | 243 | hCoV-19/USA/NY-NYUMC659/2020\|EPI_ISL_444776\|2020-0 | .................... | .................... | ..................... | .................... | .................... | .................... |
| B | 244 | hCoV-19/USA/NY-NYUMC660/2020\|EPI_ISL_444777\|2020-0 | .................... | .................... | ..................... | .................... | .................... | .................... |
| B | 245 | hCoV-19/USA/NJ-NYUMC663/2020\|EPI_ISL_444780\|2020-0 | .................... | .................... | ..................... | .................... | .................... | .................... |
| B | 246 | hCoV-19/USA/NY-NYUMC664/2020\|EPI_ISL_444781\|2020-0 | .................... | .................... | ..................... | .................... | .................... | .................... |
| B | 247 | hCoV-19/USA/NY-NYUMC665/2020\|EPI_ISL_444782\|2020-0 | .................... | .................... | ..................... | .................... | .................... | .................... |
| B | 248 | hCoV-19/USA/NY-NYUMC667/2020\|EPI_ISL_444784\|2020-0 | .................... | .................... | ..................... | .................... | .................... | .................... |
| B | 249 | hCoV-19/USA/NY-NYUMC669/2020\|EPI_ISL_444786\|2020-0 | .................... | .................... | ..................... | .................... | .................... | .................... |
| B | 250 | hCoV-19/USA/NY-NYUMC670/2020\|EPI_ISL_444787\|2020-0 | .................... | .................... | ..................... | .................... | .................... | .................... |
| B | 251 | hCoV-19/USA/NY-NYUMC671/2020\|EPI_ISL_444788\|2020-0 | .................... | .................... | ..................... | .................... | .................... | .................... |
| B | 252 | hCoV-19/USA/NY-NYUMC672/2020\|EPI_ISL_444789\|2020-0 | .................... | .................... | ..................... | .................... | .................... | .................... |
| B | 253 | hCoV-19/USA/NY-NYUMC673/2020\|EPI_ISL_444790\|2020-0 | .................... | .................... | ..................... | .................... | .................... | .................... |
| B | 254 | hCoV-19/USA/NY-NYUMC674/2020\|EPI_ISL_444791\|2020-0 | .................... | .................... | ..................... | .................... | .................... | .................... |
| B | 255 | hCoV-19/Australia/QLDID939/2020\|EPI_ISL_444793\|202 | .................... | .................... | ..................... | .................... | .................... | .................... |
| B | 256 | hCoV-19/Australia/QLDID940/2020\|EPI_ISL_444794\|202 | .................... | .................... | ..................... | .................... | .................... | .................... |
| B | 257 | hCoV-19/USA/WA-S199/2020\|EPI_ISL_430237\|2020-03-20 | .................... | .................... | ..................... | .................... | .................... | .................... |
| B | 258 | hCoV-19/England/SHEF-BFE8E/2020\|EPI_ISL_432712\|202 | .................... | .................... | ..................... | .................... | .................... | .................... |
| B | 259 | hCoV-19/England/SHEF-BFEF7/2020\|EPI_ISL_432713\|202 | .................... | .................... | ..................... | .................... | .................... | .................... |
| B | 260 | hCoV-19/England/SHEF-C0C35/2020\|EPI_ISL_432716\|202 | .................... | .................... | ..................... | .................... | .................... | .................... |
| B | 261 | hCoV-19/England/SHEF-C0FB4/2020\|EPI_ISL_432720\|202 | .................... | .................... | ..................... | .................... | .................... | .................... |
| B | 262 | hCoV-19/England/SHEF-C11BD/2020\|EPI_ISL_432722\|202 | .................... | .................... | ..................... | .................... | .................... | .................... |
| B | 263 | hCoV-19/England/SHEF-C151B/2020\|EPI_ISL_432724\|202 | .................... | .................... | ..................... | .................... | .................... | .................... |
| B | 264 | hCoV-19/England/SHEF-C1645/2020\|EPI_ISL_432727\|202 | .................... | .................... | ..................... | .................... | .................... | .................... |
| B | 266 | hCoV-19/Spain/VC-FISABIO-106/2020\|EPI_ISL_436201\|2 | .................... | .................... | ..................... | .................... | .................... | .................... |
| B | 268 | hCoV-19/Spain/VC-FISABIO-108/2020\|EPI_ISL_436203\|2 | .................... | .................... | ..................... | .................... | .................... | .................... |
| B | 269 | hCoV-19/Spain/VC-FISABIO-110/2020\|EPI_ISL_436205\|2 | .................... | .................... | ..................... | .................... | .................... | .................... |
| B | 273 | hCoV-19/Spain/VC-FISABIO-115/2020\|EPI_ISL_436210\|2 | .................... | .................... | ..................... | .................... | .................... | .................... |
| B | 274 | hCoV-19/Spain/VC-FISABIO-116/2020\|EPI_ISL_436211\|2 | .................... | .................... | ..................... | .................... | .................... | .................... |
| B | 275 | hCoV-19/Spain/VC-FISABIO-117/2020\|EPI_ISL_436212\|2 | .................... | .................... | ..................... | .................... | .................... | .................... |
| B | 276 | hCoV-19/Spain/VC-FISABIO-118/2020\|EPI_ISL_436213\|2 | .................... | .................... | ..................... | .................... | .................... | .................... |
| B | 277 | hCoV-19/Spain/VC-FISABIO-119/2020\|EPI_ISL_436214\|2 | .................... | .................... | ..................... | .................... | .................... | .................... |
| B | 279 | hCoV-19/Spain/VC-FISABIO-124/2020\|EPI_ISL_436219\|2 | .................... | .................... | ..................... | .................... | .................... | .................... |
| B | 280 | hCoV-19/Spain/VC-FISABIO-125/2020\|EPI_ISL_436220\|2 | .................... | .................... | ..................... | .................... | .................... | .................... |
| B | 281 | hCoV-19/Spain/VC-FISABIO-128/2020\|EPI_ISL_436223\|2 | .................... | .................... | ..................... | .................... | .................... | .................... |
| B | 282 | hCoV-19/Spain/VC-FISABIO-129/2020\|EPI_ISL_436224\|2 | .................... | .................... | ..................... | .................... | .................... | .................... |
| B | 283 | hCoV-19/Spain/VC-FISABIO-130/2020\|EPI_ISL_436225\|2 | .................... | .................... | ..................... | .................... | .................... | .................... |
| B | 284 | hCoV-19/Spain/VC-FISABIO-132/2020\|EPI_ISL_436227\|2 | .................... | .................... | ..................... | .................... | .................... | .................... |
| B | 286 | hCoV-19/Spain/VC-FISABIO-135/2020\|EPI_ISL_436230\|2 | .................... | .................... | ..................... | .................... | .................... | .................... |
| B | 287 | hCoV-19/Spain/VC-FISABIO-136/2020\|EPI_ISL_436231\|2 | .................... | .................... | ..................... | .................... | .................... | .................... |
| B | 291 | hCoV-19/Spain/VC-FISABIO-141/2020\|EPI_ISL_436236\|2 | .................... | .................... | ..................... | .................... | .................... | .................... |
| B | 293 | hCoV-19/Spain/VC-FISABIO-143/2020\|EPI_ISL_436238\|2 | .................... | .................... | ..................... | .................... | .................... | .................... |
| B | 295 | hCoV-19/Spain/VC-FISABIO-145/2020\|EPI_ISL_436240\|2 | .................... | .................... | ..................... | .................... | .................... | .................... |
| B | 296 | hCoV-19/Spain/VC-FISABIO-146/2020\|EPI_ISL_436241\|2 | .................... | .................... | ..................... | .................... | .................... | .................... |
| B | 297 | hCoV-19/Spain/VC-FISABIO-147/2020\|EPI_ISL_436242\|2 | .................... | .................... | ..................... | .................... | .................... | .................... |
| B | 299 | hCoV-19/Spain/VC-FISABIO-150/2020\|EPI_ISL_436245\|2 | .................... | .................... | ..................... | .................... | .................... | .................... |
| B | 300 | hCoV-19/Spain/VC-FISABIO-151/2020\|EPI_ISL_436246\|2 | .................... | .................... | ..................... | .................... | .................... | .................... |
| B | 301 | hCoV-19/Spain/VC-FISABIO-152/2020\|EPI_ISL_436247\|2 | .................... | .................... | ..................... | .................... | .................... | .................... |
| B | 302 | hCoV-19/Spain/VC-FISABIO-153/2020\|EPI_ISL_436248\|2 | .................... | .................... | ..................... | .................... | .................... | .................... |
| B | 303 | hCoV-19/Spain/VC-FISABIO-154/2020\|EPI_ISL_436249\|2 | .................... | .................... | ..................... | .................... | .................... | .................... |
| B | 304 | hCoV-19/Spain/VC-FISABIO-155/2020\|EPI_ISL_436250\|2 | .................... | .................... | ..................... | .................... | .................... | .................... |
| B | 305 | hCoV-19/Spain/VC-FISABIO-156/2020\|EPI_ISL_436251\|2 | .................... | .................... | ..................... | .................... | .................... | .................... |
| B | 306 | hCoV-19/Spain/VC-FISABIO-157/2020\|EPI_ISL_436252\|2 | .................... | .................... | ..................... | .................... | .................... | .................... |
| B | 307 | hCoV-19/Spain/VC-FISABIO-159/2020\|EPI_ISL_436254\|2 | .................... | .................... | ..................... | .................... | .................... | .................... |
| B | 308 | hCoV-19/Spain/VC-FISABIO-160/2020\|EPI_ISL_436255\|2 | .................... | .................... | ..................... | .................... | .................... | .................... |
| B | 309 | hCoV-19/Spain/VC-FISABIO-163/2020\|EPI_ISL_436258\|2 | .................... | .................... | ..................... | .................... | .................... | .................... |
| B | 311 | hCoV-19/Spain/VC-FISABIO-167/2020\|EPI_ISL_436262\|2 | .................... | .................... | ..................... | .................... | .................... | .................... |
| B | 314 | hCoV-19/Spain/VC-FISABIO-170/2020\|EPI_ISL_436265\|2 | .................... | .................... | ..................... | .................... | .................... | .................... |
| B | 317 | hCoV-19/Spain/VC-FISABIO-173/2020\|EPI_ISL_436268\|2 | .................... | .................... | ..................... | .................... | .................... | .................... |
| B | 318 | hCoV-19/Spain/VC-FISABIO-174/2020\|EPI_ISL_436269\|2 | .................... | .................... | ..................... | .................... | .................... | .................... |
| B | 319 | hCoV-19/Spain/VC-FISABIO-176/2020\|EPI_ISL_436271\|2 | .................... | .................... | ..................... | .................... | .................... | .................... |
| B | 320 | hCoV-19/Spain/VC-FISABIO-177/2020\|EPI_ISL_436272\|2 | .................... | .................... | ..................... | .................... | .................... | .................... |
| B | 321 | hCoV-19/Iran/HGRC-2-2162/2020\|EPI_ISL_437512\|2020- | .................... | .................... | ..................... | .................... | .................... | .................... |
| B | 322 | hCoV-19/USA/AK-PHL034/2020\|EPI_ISL_437513\|2020-03- | .................... | .................... | ..................... | .................... | .................... | .................... |
| B | 323 | hCoV-19/USA/AK-PHL041/2020\|EPI_ISL_437514\|2020-03- | .................... | ..........A......... | ..................... | .................... | .................... | .................... |

| | | | | | | | | |
|---|---|---|---|---|---|---|---|---|
| B | 324 hCoV-19/USA/AK-PHL070/2020\|EPI_ISL_437515\|2020-03- | ···················· | ···················· | ···················· | ···················· | ···················· | ···················· | |
| B | 325 hCoV-19/USA/AK-PHL122/2020\|EPI_ISL_437516\|2020-03- | ···················· | ···················· | ···················· | ···················· | ···················· | ···················· | |
| B | 326 hCoV-19/USA/AK-PHL178/2020\|EPI_ISL_437517\|2020-04- | ···················· | ···················· | ···················· | ···················· | ···················· | ···················· | |
| B | 327 hCoV-19/USA/AK-PHL283/2020\|EPI_ISL_437518\|2020-04- | ···················· | ···················· | ···················· | ···················· | ···················· | ···················· | |
| B | 328 hCoV-19/Czech Republic/NRL_2615/2020\|EPI_ISL_43751 | ···················· | ···················· | ···················· | ···················· | ···················· | ···················· | |
| B | 329 hCoV-19/USA/OR-OHSU-0012/2020\|EPI_ISL_437521\|2020- | ···················· | ···················· | ···················· | ···················· | ···················· | ···················· | |
| B | 330 hCoV-19/USA/OR-OHSU-0023/2020\|EPI_ISL_437523\|2020- | ···················· | ···················· | ···················· | ···················· | ···················· | ···················· | |
| B | 331 hCoV-19/USA/OR-OHSU-0027/2020\|EPI_ISL_437524\|2020- | ···················· | ···················· | ···················· | ···················· | ···················· | ···················· | |
| B | 332 hCoV-19/USA/OR-OHSU-0043/2020\|EPI_ISL_437525\|2020- | ···················· | ···················· | ···················· | ···················· | ···················· | ···················· | |
| B | 333 hCoV-19/USA/OR-OHSU-0048/2020\|EPI_ISL_437528\|2020- | ···················· | ···················· | ···················· | ···················· | ···················· | ···················· | |
| B | 334 hCoV-19/USA/OR-OHSU-0005/2020\|EPI_ISL_437529\|2020- | ···················· | ···················· | ···················· | ···················· | ···················· | ···················· | |
| B | 335 hCoV-19/USA/OR-OHSU-0051/2020\|EPI_ISL_437531\|2020- | ···················· | ···················· | ···················· | ···················· | ···················· | ···················· | |
| B | 336 hCoV-19/USA/OR-OHSU-0052/2020\|EPI_ISL_437532\|2020- | ···················· | ···················· | ···················· | ···················· | ···················· | ···················· | |
| B | 337 hCoV-19/USA/OR-OHSU-0008/2020\|EPI_ISL_437535\|2020- | ···················· | ···················· | ···················· | ···················· | ···················· | ···················· | |
| B | 338 hCoV-19/India/WB-S12/2020\|EPI_ISL_437539\|2020-04-0 | ···················· | ···················· | ···················· | ···················· | ···················· | ···················· | |
| B | 339 hCoV-19/USA/LA-SR0215/2020\|EPI_ISL_437541\|2020-04- | ···················· | ···················· | ···················· | ···················· | ···················· | ···················· | |
| B | 340 hCoV-19/USA/LA-SR0214/2020\|EPI_ISL_437542\|2020-04- | ···················· | ···················· | ···················· | ···················· | ···················· | ···················· | |
| B | 341 hCoV-19/USA/LA-SR0209/2020\|EPI_ISL_437546\|2020-03- | ···················· | ···················· | ···················· | ···················· | ···················· | ···················· | |
| B | 342 hCoV-19/USA/LA-SR0207/2020\|EPI_ISL_437547\|2020-03- | ···················· | ···················· | ···················· | ···················· | ···················· | ···················· | |
| B | 343 hCoV-19/USA/CA-SR0263/2020\|EPI_ISL_437549\|2020-04- | ···················· | ···················· | ···················· | ···················· | ···················· | ···················· | |
| B | 344 hCoV-19/USA/CA-SR0262/2020\|EPI_ISL_437550\|2020-04- | ···················· | ···················· | ···················· | ···················· | ···················· | ···················· | |
| B | 345 hCoV-19/USA/CA-SR0261/2020\|EPI_ISL_437551\|2020-04- | ···················· | ···················· | ···················· | ···················· | ···················· | ···················· | |
| B | 346 hCoV-19/USA/CA-SR0260/2020\|EPI_ISL_437552\|2020-03- | ···················· | ···················· | ···················· | ···················· | ···················· | ···················· | |
| B | 347 hCoV-19/USA/CA-SR0259/2020\|EPI_ISL_437553\|2020-03- | ···················· | ···················· | ···················· | ···················· | ···················· | ···················· | |
| B | 348 hCoV-19/USA/CA-SR0258/2020\|EPI_ISL_437554\|2020-03- | ···················· | ···················· | ···················· | ···················· | ···················· | ···················· | |
| B | 349 hCoV-19/USA/CA-SR0257/2020\|EPI_ISL_437555\|2020-03- | ···················· | ···················· | ···················· | ···················· | ···················· | ···················· | |
| B | 350 hCoV-19/USA/CA-SR0256/2020\|EPI_ISL_437556\|2020-03- | ···················· | ···················· | ···················· | ···················· | ···················· | ···················· | |
| B | 351 hCoV-19/USA/CA-SR0252/2020\|EPI_ISL_437559\|2020-03- | ···················· | ···················· | ···················· | ···················· | ···················· | ···················· | |
| B | 352 hCoV-19/USA/CA-SR0250/2020\|EPI_ISL_437560\|2020-03- | ···················· | ···················· | ···················· | ···················· | ···················· | ···················· | |
| B | 353 hCoV-19/USA/CA-SR0249/2020\|EPI_ISL_437561\|2020-03- | ···················· | ···················· | ···················· | ···················· | ···················· | ···················· | |
| B | 354 hCoV-19/USA/CA-SR0248/2020\|EPI_ISL_437562\|2020-03- | ···················· | ···················· | ···················· | ···················· | ···················· | ···················· | |
| B | 355 hCoV-19/USA/CA-SR0247/2020\|EPI_ISL_437563\|2020-03- | ···················· | ···················· | ···················· | ···················· | ···················· | ···················· | |
| B | 356 hCoV-19/USA/CA-SR0245/2020\|EPI_ISL_437564\|2020-03- | ···················· | ···················· | ···················· | ···················· | ···················· | ···················· | |
| B | 357 hCoV-19/USA/CA-SR0240/2020\|EPI_ISL_437566\|2020-03- | ···················· | ···················· | ···················· | ···················· | ···················· | ···················· | |
| B | 358 hCoV-19/USA/CA-SR0237/2020\|EPI_ISL_437567\|2020-04- | ···················· | ···················· | ···················· | ···················· | ···················· | ···················· | |
| B | 359 hCoV-19/USA/CA-SR0236/2020\|EPI_ISL_437568\|2020-04- | ···················· | ·A·················· | ···················· | ···················· | ···················· | ···················· | |
| B | 360 hCoV-19/USA/CA-SR0235/2020\|EPI_ISL_437569\|2020-04- | ···················· | ···················· | ···················· | ···················· | ···················· | ···················· | |
| B | 361 hCoV-19/USA/CA-SR0234/2020\|EPI_ISL_437570\|2020-04- | ···················· | ···················· | ···················· | ···················· | ···················· | ···················· | |
| B | 362 hCoV-19/USA/CA-SR0233/2020\|EPI_ISL_437571\|2020-04- | ···················· | ·A·················· | ···················· | ···················· | ···················· | ···················· | |
| B | 363 hCoV-19/USA/CA-SR0232/2020\|EPI_ISL_437572\|2020-04- | ···················· | ·A·················· | ···················· | ···················· | ···················· | ···················· | |
| B | 364 hCoV-19/USA/CA-SR0228/2020\|EPI_ISL_437574\|2020-04- | ···················· | ···················· | ···················· | ···················· | ···················· | ···················· | |
| B | 365 hCoV-19/USA/CA-SR0225/2020\|EPI_ISL_437575\|2020-04- | ···················· | ·A·················· | ···················· | ···················· | ···················· | ···················· | |
| B | 366 hCoV-19/USA/CA-SR0224/2020\|EPI_ISL_437576\|2020-04- | ···················· | ···················· | ···················· | ···················· | ···················· | ···················· | |
| B | 367 hCoV-19/USA/CA-SR0218/2020\|EPI_ISL_437580\|2020-04- | ···················· | ·A·················· | ···················· | ···················· | ···················· | ···················· | |
| B | 368 hCoV-19/USA/CA-SR0204/2020\|EPI_ISL_437582\|2020-03- | ···················· | ···················· | ···················· | ···················· | ···················· | ···················· | |
| B | 369 hCoV-19/USA/CA-SR0201/2020\|EPI_ISL_437583\|2020-03- | ···················· | ···················· | ···················· | ···················· | ···················· | ···················· | |
| B | 370 hCoV-19/USA/CA-SR0199/2020\|EPI_ISL_437584\|2020-03- | ···················· | ···················· | ···················· | ···················· | ···················· | ···················· | |
| B | 371 hCoV-19/USA/CA-SR0198/2020\|EPI_ISL_437585\|2020-03- | ···················· | ···················· | ···················· | ···················· | ···················· | ···················· | |
| B | 372 hCoV-19/USA/CA-SR0197/2020\|EPI_ISL_437586\|2020-03- | ···················· | ···················· | ···················· | ···················· | ···················· | ···················· | |
| B | 373 hCoV-19/USA/CA-SR0195/2020\|EPI_ISL_437587\|2020-03- | ···················· | ···················· | ···················· | ···················· | ···················· | ···················· | |
| B | 374 hCoV-19/USA/CA-SR0193/2020\|EPI_ISL_437588\|2020-03- | ···················· | ···················· | ···················· | ···················· | ···················· | ···················· | |
| B | 375 hCoV-19/USA/CA-SR0192/2020\|EPI_ISL_437589\|2020-03- | ···················· | ···················· | ···················· | ···················· | ···················· | ···················· | |
| B | 376 hCoV-19/USA/CA-SR0190/2020\|EPI_ISL_437590\|2020-03- | ···················· | ···················· | ···················· | ···················· | ···················· | ···················· | |
| B | 377 hCoV-19/USA/CA-SR0189/2020\|EPI_ISL_437591\|2020-03- | ···················· | ···················· | ···················· | ···················· | ···················· | ···················· | |
| B | 378 hCoV-19/USA/CA-SR0188/2020\|EPI_ISL_437592\|2020-03- | ···················· | ···················· | ···················· | ···················· | ···················· | ···················· | |
| B | 379 hCoV-19/USA/CA-SR0184/2020\|EPI_ISL_437594\|2020-03- | ···················· | ···················· | ···················· | ···················· | ···················· | ···················· | |
| B | 380 hCoV-19/USA/CA-SR0182/2020\|EPI_ISL_437595\|2020-03- | ···················· | ···················· | ···················· | ···················· | ···················· | ···················· | |
| B | 381 hCoV-19/USA/CA-SR0181/2020\|EPI_ISL_437596\|2020-03- | ···················· | ···················· | ···················· | ···················· | ···················· | ···················· | |
| B | 382 hCoV-19/USA/CA-SR0179/2020\|EPI_ISL_437598\|2020-03- | ···················· | ···················· | ···················· | ···················· | ···················· | ···················· | |
| B | 383 hCoV-19/USA/CA-SR0178/2020\|EPI_ISL_437599\|2020-03- | ···················· | ···················· | ···················· | ···················· | ···················· | ···················· | |
| B | 384 hCoV-19/Scotland/CVR1427/2020\|EPI_ISL_438806\|2020- | ···················· | ···················A | ···················· | ···················· | ···················· | ···················· | 1 |
| B | 385 hCoV-19/Scotland/CVR1448/2020\|EPI_ISL_438815\|2020- | ···················· | ···················· | ···················· | ···················· | ···················· | ···················· | |
| B | 386 hCoV-19/Scotland/CVR1484/2020\|EPI_ISL_438824\|2020- | ···················· | ···················· | ···················· | ···················· | ···················· | ···················· | |
| B | 387 hCoV-19/Scotland/CVR1496/2020\|EPI_ISL_438827\|2020- | ···················· | ·A·················· | ···················· | ···················· | ···················· | ···················· | |
| B | 388 hCoV-19/Scotland/CVR1499/2020\|EPI_ISL_438829\|2020- | ···················· | ···················· | ···················· | ···················· | ···················· | ···················· | |
| B | 389 hCoV-19/Scotland/CVR1504/2020\|EPI_ISL_438830\|2020- | ···················· | ···················A | ···················· | ···················· | ···················· | ···················· | 1 |
| B | 390 hCoV-19/Scotland/CVR1506/2020\|EPI_ISL_438831\|2020- | ···················· | ···················A | ···················· | ···················· | ···················· | ···················· | 1 |
| B | 391 hCoV-19/Scotland/CVR1533/2020\|EPI_ISL_438836\|2020- | ···················· | ···················A | ···················· | ···················· | ···················· | ···················· | 1 |
| B | 392 hCoV-19/Scotland/CVR1537/2020\|EPI_ISL_438837\|2020- | ···················· | ···················· | ···················· | ···················· | ···················· | ···················· | |
| B | 393 hCoV-19/Scotland/CVR1557/2020\|EPI_ISL_438839\|2020- | ···················· | ···················· | ···················· | ···················· | ···················· | ···················· | |
| B | 394 hCoV-19/Scotland/CVR1560/2020\|EPI_ISL_438840\|2020- | ···················· | ···················· | ···················· | ···················· | ···················· | ···················· | |
| B | 395 hCoV-19/Scotland/CVR1573/2020\|EPI_ISL_438841\|2020- | ···················· | ···················· | ···················· | ···················· | ···················· | ···················· | |
| B | 396 hCoV-19/Scotland/CVR1579/2020\|EPI_ISL_438842\|2020- | ···················· | ···················· | ···················· | ···················· | ···················· | ···················· | |
| B | 397 hCoV-19/Scotland/CVR1615/2020\|EPI_ISL_438844\|2020- | ···················· | ···················· | ···················· | ···················· | ···················· | ···················· | |
| B | 399 hCoV-19/Scotland/CVR1816/2020\|EPI_ISL_438849\|2020- | ···················· | ···················· | ···················· | ···················· | ···················· | ···················· | |
| B | 400 hCoV-19/Scotland/CVR1843/2020\|EPI_ISL_438850\|2020- | ···················· | ···················· | ···················· | ···················· | ···················· | ···················· | |
| B | 401 hCoV-19/Scotland/CVR1951/2020\|EPI_ISL_438860\|2020- | ···················· | ·A·················· | ···················· | ···················· | ···················· | ···················· | |
| B | 402 hCoV-19/Scotland/CVR1997/2020\|EPI_ISL_438868\|2020- | ···················· | ·A·················· | ···················· | ···················· | ···················· | ···················· | |

| | | | | | | | | | |
|---|---|---|---|---|---|---|---|---|---|
| B | 403 | hCoV-19/Scotland/CVR2012/2020\|EPI_ISL_438870\|2020- | .................... | .................... | .................... | .................... | .................... | .................... | |
| B | 404 | hCoV-19/Scotland/CVR2124/2020\|EPI_ISL_438875\|2020- | .................... | .................... | .................... | .................... | .................... | .................... | |
| B | 405 | hCoV-19/Scotland/CVR2153/2020\|EPI_ISL_438877\|2020- | .................... | .................... | .................... | .................... | .................... | .................... | |
| B | 406 | hCoV-19/Scotland/CVR2179/2020\|EPI_ISL_438878\|2020- | .................... | .................... | .................... | .................... | .................... | .................... | |
| B | 407 | hCoV-19/Scotland/CVR2215/2020\|EPI_ISL_438879\|2020- | .................... | .................... | .................... | .................... | .................... | .................... | |
| B | 408 | hCoV-19/Scotland/CVR226/2020\|EPI_ISL_438883\|2020-0 | .................... | .................... | .................... | .................... | .................... | .................... | |
| B | 409 | hCoV-19/Scotland/CVR2260/2020\|EPI_ISL_438884\|2020- | .................... | .................... | .................... | .................... | .................... | .................... | |
| B | 410 | hCoV-19/Scotland/CVR234/2020\|EPI_ISL_438889\|2020-0 | .................... | .................... | .................... | .................... | .................... | .................... | |
| B | 411 | hCoV-19/Scotland/CVR2347/2020\|EPI_ISL_438890\|2020- | .................... | .................... | .................... | .................... | .................... | .................... | |
| B | 412 | hCoV-19/Scotland/CVR2349/2020\|EPI_ISL_438891\|2020- | .................... | .................... | .................... | .................... | .................... | .................... | |
| B | 413 | hCoV-19/Scotland/CVR2357/2020\|EPI_ISL_438892\|2020- | .................... | .................... | .................... | .................... | .................... | .................... | |
| B | 414 | hCoV-19/Scotland/CVR2367/2020\|EPI_ISL_438893\|2020- | .................... | ...................A | .................... | .................... | .................... | .................... | 1 |
| B | 415 | hCoV-19/Scotland/CVR2372/2020\|EPI_ISL_438894\|2020- | .................... | .................... | .................... | .................... | .................... | .................... | |
| B | 416 | hCoV-19/Scotland/CVR2378/2020\|EPI_ISL_438895\|2020- | .................... | .................... | .................... | .................... | .................... | .................... | |
| B | 417 | hCoV-19/Scotland/CVR238/2020\|EPI_ISL_438896\|2020-0 | .................... | .................... | .................... | .................... | .................... | .................... | |
| B | 418 | hCoV-19/England/SHEF-C9935/2020\|EPI_ISL_442043\|202 | .................... | .................... | .................... | .................... | .................... | .................... | |
| B | 419 | hCoV-19/Iran/KHGRC-2-2162/2020\|EPI_ISL_442044\|2020 | .................... | .................... | .................... | .................... | .................... | .................... | |
| B | 420 | hCoV-19/England/CAMB-7D077/2020\|EPI_ISL_442045\|202 | .................... | .................... | .................... | .................... | .................... | .................... | |
| B | 421 | hCoV-19/England/CAMB-7BC66/2020\|EPI_ISL_442046\|202 | .................... | .................... | .................... | .................... | .................... | .................... | |
| B | 422 | hCoV-19/England/CAMB-7CBD1/2020\|EPI_ISL_442047\|202 | .................... | .................... | .................... | .................... | .................... | .................... | |
| B | 423 | hCoV-19/England/CAMB-7C7A0/2020\|EPI_ISL_442048\|202 | .................... | .................... | .................... | .................... | .................... | .................... | |
| B | 424 | hCoV-19/England/CAMB-7CA89/2020\|EPI_ISL_442049\|202 | .................... | .................... | .................... | .................... | .................... | .................... | |
| B | 425 | hCoV-19/England/CAMB-7C931/2020\|EPI_ISL_442050\|202 | .................... | .................... | .................... | .................... | .................... | .................... | |
| B | 426 | hCoV-19/England/CAMB-7D1DE/2020\|EPI_ISL_442051\|202 | .................... | .................... | .................... | .................... | .................... | .................... | |
| B | 427 | hCoV-19/England/CAMB-7BE24/2020\|EPI_ISL_442053\|202 | .................... | .................... | .................... | .................... | .................... | .................... | |
| B | 428 | hCoV-19/England/CAMB-7C8AD/2020\|EPI_ISL_442054\|202 | .................... | .................... | .................... | .................... | .................... | .................... | |
| B | 429 | hCoV-19/England/CAMB-7C7CE/2020\|EPI_ISL_442055\|202 | .................... | .................... | .................... | .................... | .................... | .................... | |
| B | 430 | hCoV-19/England/CAMB-7E0FE/2020\|EPI_ISL_442056\|202 | .................... | .................... | .................... | .................... | .................... | .................... | |
| B | 431 | hCoV-19/England/CAMB-7CC56/2020\|EPI_ISL_442057\|202 | .................... | .................... | .................... | .................... | .................... | .................... | |
| B | 432 | hCoV-19/England/CAMB-7D1B0/2020\|EPI_ISL_442058\|202 | .................... | .................... | .................... | .................... | .................... | .................... | |
| B | 433 | hCoV-19/England/CAMB-7D059/2020\|EPI_ISL_442059\|202 | .................... | .................... | .................... | .................... | .................... | .................... | |
| B | 434 | hCoV-19/England/CAMB-7B9C9/2020\|EPI_ISL_442060\|202 | .................... | .................... | .................... | .................... | .................... | .................... | |
| B | 435 | hCoV-19/England/CAMB-7C870/2020\|EPI_ISL_442061\|202 | .................... | .................... | .................... | .................... | .................... | .................... | |
| B | 436 | hCoV-19/England/CAMB-7BD81/2020\|EPI_ISL_442062\|202 | .................... | .................... | .................... | .................... | .................... | .................... | |
| B | 437 | hCoV-19/England/CAMB-7CAA7/2020\|EPI_ISL_442064\|202 | .................... | .................... | .................... | .................... | .................... | .................... | |
| B | 438 | hCoV-19/England/CAMB-7B871/2020\|EPI_ISL_442066\|202 | .................... | .................... | .................... | .................... | .................... | .................... | |
| B | 439 | hCoV-19/England/CAMB-7D271/2020\|EPI_ISL_442067\|202 | .................... | .................... | .................... | .................... | .................... | .................... | |
| B | 440 | hCoV-19/England/CAMB-7C99B/2020\|EPI_ISL_442068\|202 | .................... | .................... | .................... | .................... | .................... | .................... | |
| B | 441 | hCoV-19/England/CAMB-7E01C/2020\|EPI_ISL_442070\|202 | .................... | .................... | .................... | .................... | .................... | .................... | |
| B | 442 | hCoV-19/England/CAMB-7BC39/2020\|EPI_ISL_442071\|202 | .................... | .................... | .................... | .................... | .................... | .................... | |
| B | 443 | hCoV-19/England/CAMB-7D29F/2020\|EPI_ISL_442073\|202 | .................... | .................... | .................... | .................... | .................... | .................... | |
| B | 444 | hCoV-19/England/CAMB-7C9E6/2020\|EPI_ISL_442074\|202 | .................... | .................... | .................... | .................... | .................... | .................... | |
| B | 445 | hCoV-19/England/CAMB-7BBB4/2020\|EPI_ISL_442075\|202 | .................... | .................... | .................... | .................... | .................... | .................... | |
| B | 446 | hCoV-19/England/CAMB-7C8CB/2020\|EPI_ISL_442076\|202 | .................... | .................... | .................... | .................... | .................... | .................... | |
| B | 447 | hCoV-19/England/CAMB-7C728/2020\|EPI_ISL_442077\|202 | .................... | .................... | .................... | .................... | .................... | .................... | |
| B | 448 | hCoV-19/England/CAMB-7CDF9/2020\|EPI_ISL_442078\|202 | .................... | .................... | .................... | .................... | .................... | .................... | |
| B | 449 | hCoV-19/England/CAMB-7CB1D/2020\|EPI_ISL_442079\|202 | .................... | .................... | .................... | .................... | .................... | .................... | |
| B | 450 | hCoV-19/England/CAMB-7BF7C/2020\|EPI_ISL_442080\|202 | .................... | .................... | .................... | .................... | .................... | .................... | |
| B | 451 | hCoV-19/England/CAMB-7DE40/2020\|EPI_ISL_442081\|202 | .................... | .................... | .................... | .................... | .................... | .................... | |
| B | 452 | hCoV-19/England/CAMB-7CB3B/2020\|EPI_ISL_442082\|202 | .................... | .................... | .................... | .................... | .................... | .................... | |
| B | 453 | hCoV-19/England/CAMB-7DE6E/2020\|EPI_ISL_442083\|202 | .................... | .................... | .................... | .................... | .................... | .................... | |
| B | 454 | hCoV-19/England/CAMB-7DD43/2020\|EPI_ISL_442084\|202 | .................... | .................... | .................... | .................... | .................... | .................... | |
| B | 455 | hCoV-19/England/CAMB-7CA10/2020\|EPI_ISL_442085\|202 | .................... | .................... | .................... | .................... | .................... | .................... | |
| B | 456 | hCoV-19/England/CAMB-7BE06/2020\|EPI_ISL_442086\|202 | .................... | .................... | .................... | .................... | .................... | .................... | |
| B | 457 | hCoV-19/England/CAMB-7CAF2/2020\|EPI_ISL_442088\|202 | .................... | .................... | .................... | .................... | .................... | .................... | |
| B | 458 | hCoV-19/England/CAMB-7BC0C/2020\|EPI_ISL_442089\|202 | .................... | .................... | .................... | .................... | .................... | .................... | |
| B | 459 | hCoV-19/England/CAMB-7D226/2020\|EPI_ISL_442090\|202 | .................... | .................... | .................... | .................... | .................... | .................... | |
| B | 460 | hCoV-19/England/CAMB-7CAC5/2020\|EPI_ISL_442091\|202 | .................... | .................... | .................... | .................... | .................... | .................... | |
| B | 461 | hCoV-19/England/CAMB-7DE04/2020\|EPI_ISL_442092\|202 | .................... | .................... | .................... | .................... | .................... | .................... | |
| B | 462 | hCoV-19/England/CAMB-7E049/2020\|EPI_ISL_442093\|202 | .................... | .................... | .................... | .................... | .................... | .................... | |
| B | 463 | hCoV-19/England/CAMB-7BC93/2020\|EPI_ISL_442094\|202 | .................... | .................... | .................... | .................... | .................... | .................... | |
| B | 464 | hCoV-19/England/CAMB-7D01D/2020\|EPI_ISL_442095\|202 | .................... | .................... | .................... | .................... | .................... | ...............A.... | |
| B | 465 | hCoV-19/England/CAMB-7BCEE/2020\|EPI_ISL_442097\|202 | .................... | .................... | .................... | .................... | .................... | .................... | |
| B | 466 | hCoV-19/England/CAMB-7C7EC/2020\|EPI_ISL_442098\|202 | .................... | .................... | .................... | .................... | .................... | .................... | |
| B | 467 | hCoV-19/England/CAMB-7CA7A/2020\|EPI_ISL_442220\|202 | .................... | .................... | .................... | .................... | .................... | .................... | |
| B | 468 | hCoV-19/England/CAMB-7DBEF/2020\|EPI_ISL_442221\|202 | .................... | .................... | .................... | .................... | .................... | .................... | |
| B | 469 | hCoV-19/England/CAMB-7C49A/2020\|EPI_ISL_442222\|202 | .................... | .................... | .................... | .................... | .................... | .................... | |
| B | 470 | hCoV-19/England/CAMB-7DAA6/2020\|EPI_ISL_442223\|202 | .................... | .................... | .................... | .................... | .................... | .................... | |
| B | 471 | hCoV-19/England/CAMB-83CC5/2020\|EPI_ISL_443371\|202 | .................... | .................... | .................... | .................... | .................... | .................... | |
| B | 472 | hCoV-19/England/CAMB-83CE3/2020\|EPI_ISL_443372\|202 | .................... | .................... | .................... | .................... | .................... | .................... | |
| B | 473 | hCoV-19/England/CAMB-83C10/2020\|EPI_ISL_443375\|202 | .................... | .................... | .................... | .................... | .................... | .................... | |
| B | 474 | hCoV-19/England/CAMB-83991/2020\|EPI_ISL_443378\|202 | .................... | .................... | .................... | .................... | .................... | .................... | |
| B | 475 | hCoV-19/England/CAMB-83937/2020\|EPI_ISL_443383\|202 | .................... | .................... | .................... | .................... | .................... | .................... | |
| B | 476 | hCoV-19/England/CAMB-8366D/2020\|EPI_ISL_443384\|202 | .................... | .................... | .................... | .................... | .................... | .................... | |
| B | 477 | hCoV-19/England/CAMB-8258F/2020\|EPI_ISL_443385\|202 | .................... | .................... | .................... | .................... | .................... | .................... | |
| B | 478 | hCoV-19/England/CAMB-830E7/2020\|EPI_ISL_443386\|202 | .................... | .................... | .................... | .................... | .................... | .................... | |
| B | 479 | hCoV-19/England/CAMB-82DB4/2020\|EPI_ISL_443387\|202 | .................... | .................... | .................... | .................... | .................... | .................... | |
| B | 480 | hCoV-19/England/CAMB-82D96/2020\|EPI_ISL_443388\|202 | .................... | .................... | .................... | .................... | .................... | .................... | |

| | | | | | | | | |
|---|---|---|---|---|---|---|---|---|
| B | 481 | hCoV-19/England/CAMB-82E48/2020\|EPI_ISL_443389\|202 | .................... | .................... | ..................... | .................... | .................... | .................... |
| B | 482 | hCoV-19/England/CAMB-832E1/2020\|EPI_ISL_443390\|202 | .................... | .................... | ..................... | .................... | .................... | .................... |
| B | 483 | hCoV-19/England/CAMB-83867/2020\|EPI_ISL_443391\|202 | .................... | .................... | ..................... | .................... | .................... | .................... |
| B | 484 | hCoV-19/England/CAMB-82622/2020\|EPI_ISL_443392\|202 | .................... | .................... | ..................... | .................... | .................... | .................... |
| B | 485 | hCoV-19/England/CAMB-83603/2020\|EPI_ISL_443393\|202 | .................... | .................... | ..................... | .................... | .................... | .................... |
| B | 486 | hCoV-19/England/CAMB-835CA/2020\|EPI_ISL_443394\|202 | .................... | .................... | ..................... | .................... | .................... | .................... |
| B | 487 | hCoV-19/England/CAMB-834CD/2020\|EPI_ISL_443395\|202 | .................... | .................... | ..................... | .................... | .................... | .................... |
| B | 488 | hCoV-19/England/CAMB-832C3/2020\|EPI_ISL_443396\|202 | .................... | .................... | ..................... | .................... | .................... | .................... |
| B | 489 | hCoV-19/England/CAMB-8320F/2020\|EPI_ISL_443398\|202 | .................... | .................... | ..................... | .................... | .................... | .................... |
| B | 490 | hCoV-19/England/CAMB-82604/2020\|EPI_ISL_443399\|202 | .................... | .................... | ..................... | .................... | .................... | .................... |
| B | 491 | hCoV-19/England/CAMB-82CF3/2020\|EPI_ISL_443552\|202 | .................... | .................... | ..................... | .................... | .................... | .................... |
| B | 492 | hCoV-19/England/CAMB-1AA2A7/2020\|EPI_ISL_443554\|20 | .................... | .................... | ..................... | .................... | .................... | .................... |
| B | 493 | hCoV-19/USA/NY-NYUMC582/2020\|EPI_ISL_444699\|2020-0 | .................... | .................... | ..................... | .................... | .................... | .................... |
| B | 494 | hCoV-19/USA/NY-NYUMC583/2020\|EPI_ISL_444700\|2020-0 | .................... | .................... | ..................... | .................... | .................... | .................... |
| B | 495 | hCoV-19/USA/NY-NYUMC584/2020\|EPI_ISL_444701\|2020-0 | .................... | .................... | ..................... | .................... | .................... | .................... |
| B | 496 | hCoV-19/USA/NY-NYUMC585/2020\|EPI_ISL_444702\|2020-0 | .................... | .................... | ..................... | .................... | .................... | .................... |
| B | 497 | hCoV-19/USA/NY-NYUMC586/2020\|EPI_ISL_444703\|2020-0 | .................... | .................... | ..................... | .................... | .................... | .................... |
| B | 498 | hCoV-19/USA/NY-NYUMC587/2020\|EPI_ISL_444704\|2020-0 | .................... | .................... | ..................... | .................... | .................... | .................... |
| B | 499 | hCoV-19/USA/NY-NYUMC588/2020\|EPI_ISL_444705\|2020-0 | .................... | .................... | ..................... | .................... | .................... | .................... |
| B | 500 | hCoV-19/USA/NY-NYUMC589/2020\|EPI_ISL_444706\|2020-0 | .................... | .................... | ..................... | .................... | .................... | .................... |
| B | 501 | hCoV-19/USA/NY-NYUMC590/2020\|EPI_ISL_444707\|2020-0 | .................... | .................... | ..................... | .................... | .................... | .................... |
| B | 502 | hCoV-19/USA/NY-NYUMC591/2020\|EPI_ISL_444708\|2020-0 | .................... | .................... | ..................... | .................... | .................... | .................... |
| B | 503 | hCoV-19/USA/NY-NYUMC604/2020\|EPI_ISL_444721\|2020-0 | .................... | .................... | ..................... | .................... | .................... | .................... |
| B | 504 | hCoV-19/USA/NY-NYUMC608/2020\|EPI_ISL_444725\|2020-0 | .................... | .................... | ..................... | .................... | .................... | .................... |
| B | 505 | hCoV-19/USA/NY-NYUMC609/2020\|EPI_ISL_444726\|2020-0 | .................... | .................... | ..................... | .................... | .................... | .................... |
| B | 506 | hCoV-19/USA/NY-NYUMC610/2020\|EPI_ISL_444727\|2020-0 | .................... | .................... | ..................... | .................... | .................... | .................... |
| B | 507 | hCoV-19/USA/NY-NYUMC611/2020\|EPI_ISL_444728\|2020-0 | .................... | .................... | ..................... | .................... | .................... | .................... |
| B | 508 | hCoV-19/Denmark/ALAB-SSI-731/2020\|EPI_ISL_444880\|2 | .................... | .................... | ..................... | .................... | .................... | .................... |
| B | 509 | hCoV-19/Denmark/ALAB-SSI-735/2020\|EPI_ISL_444881\|2 | .................... | .................... | ..................... | .................... | .................... | .................... |
| B | 510 | hCoV-19/Denmark/ALAB-SSI-741/2020\|EPI_ISL_444883\|2 | .................... | .................... | ..................... | .................... | .................... | .................... |
| B | 511 | hCoV-19/Denmark/ALAB-SSI-742/2020\|EPI_ISL_444884\|2 | .................... | .................... | ..................... | .................... | .................... | .................... |
| B | 512 | hCoV-19/Denmark/ALAB-SSI-743/2020\|EPI_ISL_444885\|2 | .................... | .................... | ..................... | .................... | .................... | .................... |
| B | 513 | hCoV-19/USA/WA-S174/2020\|EPI_ISL_430212\|2020-03-19 | .................... | .................... | ..................... | .................... | .................... | .................... |
| B | 514 | hCoV-19/USA/WA-S175/2020\|EPI_ISL_430213\|2020-03-19 | .................... | .................... | ..................... | .................... | .................... | .................... |
| B | 515 | hCoV-19/USA/WA-S176/2020\|EPI_ISL_430214\|2020-03-19 | .................... | .................... | ..................... | .................... | .................... | .................... |
| B | 516 | hCoV-19/USA/WA-S177/2020\|EPI_ISL_430215\|2020-03-19 | .................... | .................... | ..................... | .................... | .................... | .................... |
| B | 517 | hCoV-19/USA/WA-S178/2020\|EPI_ISL_430216\|2020-03-20 | .................... | .................... | ..................... | .................... | .................... | .................... |
| B | 518 | hCoV-19/USA/WA-S179/2020\|EPI_ISL_430217\|2020-03-20 | .................... | .................... | ..................... | .................... | .................... | .................... |
| B | 519 | hCoV-19/USA/WA-S181/2020\|EPI_ISL_430219\|2020-03-19 | .................... | .................... | ..................... | .................... | .................... | .................... |
| B | 520 | hCoV-19/USA/WA-S182/2020\|EPI_ISL_430220\|2020-03-20 | .................... | .................... | ..................... | .................... | .................... | .................... |
| B | 521 | hCoV-19/USA/WA-S183/2020\|EPI_ISL_430221\|2020-03-19 | .................... | .................... | ..................... | .................... | .................... | .................... |
| B | 522 | hCoV-19/USA/WA-S184/2020\|EPI_ISL_430222\|2020-03-19 | .................... | .................... | ..................... | .................... | .................... | .................... |
| B | 523 | hCoV-19/USA/WA-S185/2020\|EPI_ISL_430223\|2020-03-19 | .................... | .................... | ..................... | .................... | .................... | .................... |
| B | 524 | hCoV-19/USA/WA-S186/2020\|EPI_ISL_430224\|2020-03-20 | .................... | .................... | ..................... | .................... | .................... | .................... |
| B | 525 | hCoV-19/USA/WA-S187/2020\|EPI_ISL_430225\|2020-03-20 | .................... | .................... | ..................... | .................... | .................... | .................... |
| B | 526 | hCoV-19/USA/WA-S188/2020\|EPI_ISL_430226\|2020-03-20 | .................... | .................... | ..................... | .................... | .................... | .................... |
| B | 527 | hCoV-19/USA/WA-S189/2020\|EPI_ISL_430227\|2020-03-20 | .................... | .................... | ..................... | .................... | .................... | .................... |
| B | 528 | hCoV-19/USA/WA-S190/2020\|EPI_ISL_430228\|2020-03-23 | .................... | .................... | ..................... | .................... | .................... | .................... |
| B | 529 | hCoV-19/USA/WA-S191/2020\|EPI_ISL_430229\|2020-03-20 | .................... | .................... | ..................... | .................... | .................... | .................... |
| B | 530 | hCoV-19/USA/WA-S192/2020\|EPI_ISL_430230\|2020-03-20 | .................... | .................... | ..................... | .................... | .................... | .................... |
| B | 531 | hCoV-19/USA/WA-S193/2020\|EPI_ISL_430231\|2020-03-19 | .................... | .................... | ..................... | .................... | .................... | .................... |
| B | 532 | hCoV-19/USA/WA-S194/2020\|EPI_ISL_430232\|2020-03-20 | .................... | .................... | ..................... | .................... | .................... | .................... |
| B | 533 | hCoV-19/USA/WA-S195/2020\|EPI_ISL_430233\|2020-03-20 | .................... | .................... | ..................... | .................... | .................... | .................... |
| B | 534 | hCoV-19/USA/WA-S196/2020\|EPI_ISL_430234\|2020-03-19 | .................... | .................... | ..................... | .................... | .................... | .................... |
| B | 535 | hCoV-19/USA/WA-S197/2020\|EPI_ISL_430235\|2020-03-20 | .................... | .................... | ..................... | .................... | .................... | .................... |
| B | 536 | hCoV-19/USA/WA-S198/2020\|EPI_ISL_430236\|2020-03-20 | .................... | .................... | ..................... | .................... | .................... | .................... |
| B | 537 | hCoV-19/USA/WA-S200/2020\|EPI_ISL_430238\|2020-03-20 | .................... | .................... | ..................... | .................... | .................... | .................... |
| B | 538 | hCoV-19/USA/WA-S201/2020\|EPI_ISL_430239\|2020-03-20 | .................... | .................... | ..................... | .................... | .................... | .................... |
| B | 539 | hCoV-19/USA/WA-S202/2020\|EPI_ISL_430240\|2020-03-20 | .................... | .................... | ..................... | .................... | .................... | .................... |
| B | 540 | hCoV-19/USA/WA-S203/2020\|EPI_ISL_430241\|2020-03-20 | .................... | .................... | ..................... | .................... | .................... | .................... |
| B | 541 | hCoV-19/USA/WA-S204/2020\|EPI_ISL_430242\|2020-03-22 | .................... | .................... | ..................... | .................... | .................... | .................... |
| B | 542 | hCoV-19/USA/WA-S205/2020\|EPI_ISL_430243\|2020-03-20 | .................... | .................... | ..................... | .................... | .................... | .................... |
| B | 543 | hCoV-19/USA/WA-S206/2020\|EPI_ISL_430244\|2020-03-20 | .................... | .................... | ..................... | .................... | .................... | .................... |
| B | 544 | hCoV-19/USA/WA-S207/2020\|EPI_ISL_430245\|2020-03-20 | .................... | .................... | ..................... | .................... | .................... | .................... |
| B | 545 | hCoV-19/USA/WA-S208/2020\|EPI_ISL_430246\|2020-03-23 | .................... | .................... | ..................... | .................... | .................... | .................... |
| B | 546 | hCoV-19/USA/WA-S209/2020\|EPI_ISL_430247\|2020-03-23 | .................... | .................... | ..................... | .................... | .................... | .................... |
| B | 547 | hCoV-19/USA/WA-S210/2020\|EPI_ISL_430248\|2020-03-19 | .................... | .................... | ..................... | .................... | .................... | .................... |
| B | 548 | hCoV-19/USA/WA-S211/2020\|EPI_ISL_430249\|2020-03-20 | .................... | .................... | ..................... | .................... | .................... | .................... |
| B | 549 | hCoV-19/USA/WA-S212/2020\|EPI_ISL_430250\|2020-03-19 | .................... | .................... | ..................... | .................... | .................... | .................... |
| B | 550 | hCoV-19/USA/WA-S214/2020\|EPI_ISL_430252\|2020-03-20 | .................... | .................... | ..................... | .................... | .................... | .................... |
| B | 551 | hCoV-19/USA/WA-S215/2020\|EPI_ISL_430253\|2020-03-20 | .................... | .................... | ..................... | .................... | .................... | .................... |
| B | 552 | hCoV-19/USA/WA-S216/2020\|EPI_ISL_430254\|2020-03-20 | .................... | .................... | ..................... | .................... | .................... | .................... |
| B | 553 | hCoV-19/USA/WA-S217/2020\|EPI_ISL_430255\|2020-03-20 | .................... | .................... | ..................... | .................... | .................... | .................... |
| B | 554 | hCoV-19/USA/WA-S218/2020\|EPI_ISL_430256\|2020-03-23 | .................... | .................... | ..................... | .................... | .................... | .................... |
| B | 555 | hCoV-19/USA/WA-S219/2020\|EPI_ISL_430257\|2020-03-20 | .................... | .................... | ..................... | .................... | .................... | .................... |
| B | 556 | hCoV-19/USA/WA-S220/2020\|EPI_ISL_430258\|2020-03-20 | .................... | .................... | ..................... | .................... | .................... | .................... |
| B | 557 | hCoV-19/USA/WA-S221/2020\|EPI_ISL_430259\|2020-03-19 | .................... | .................... | ..................... | .................... | .................... | .................... |
| B | 558 | hCoV-19/USA/WA-S223/2020\|EPI_ISL_430261\|2020-03-20 | .................... | .................... | ..................... | .................... | .................... | .................... |

| | | | | | | | |
|---|---|---|---|---|---|---|---|
| B | 559 hCoV-19/USA/WA-S224/2020\|EPI_ISL_430262\|2020-03-19 | ···················· | ···················· | ···················· | ···················· | ···················· | ···················· |
| B | 560 hCoV-19/USA/WA-S225/2020\|EPI_ISL_430263\|2020-03-20 | ···················· | ···················· | ···················· | ···················· | ···················· | ···················· |
| B | 561 hCoV-19/USA/WA-S226/2020\|EPI_ISL_430264\|2020-03-21 | ···················· | ···················· | ···················· | ···················· | ···················· | ···················· |
| B | 562 hCoV-19/USA/WA-S227/2020\|EPI_ISL_430265\|2020-03-20 | ···················· | ···················· | ···················· | ···················· | ···················· | ···················· |
| B | 563 hCoV-19/USA/WA-S228/2020\|EPI_ISL_430266\|2020-03-20 | ···················· | ···················· | ···················· | ···················· | ···················· | ···················· |
| B | 564 hCoV-19/USA/WA-S229/2020\|EPI_ISL_430267\|2020-03-19 | ···················· | ···················· | ···················· | ···················· | ···················· | ···················· |
| B | 565 hCoV-19/USA/WA-S230/2020\|EPI_ISL_430268\|2020-03-19 | ···················· | ···················· | ···················· | ···················· | ···················· | ···················· |
| B | 566 hCoV-19/USA/WA-S231/2020\|EPI_ISL_430269\|2020-03-20 | ···················· | ···················· | ···················· | ···················· | ···················· | ···················· |
| B | 567 hCoV-19/USA/WA-S232/2020\|EPI_ISL_430270\|2020-03-08 | ···················· | ···················· | ···················· | ···················· | ···················· | ···················· |
| B | 568 hCoV-19/USA/WA-S233/2020\|EPI_ISL_430271\|2020-03-03 | ···················· | ···················· | ···················· | ···················· | ···················· | ···················· |
| B | 569 hCoV-19/USA/WA-S234/2020\|EPI_ISL_430272\|2020-02-28 | ···················· | ···················· | ···················· | ···················· | ···················· | ···················· |
| B | 570 hCoV-19/USA/WA-S235/2020\|EPI_ISL_430273\|2020-02-29 | ···················· | ···················· | ···················· | ···················· | ···················· | ···················· |
| B | 571 hCoV-19/USA/WA-S236/2020\|EPI_ISL_430274\|2020-03-01 | ···················· | ···················· | ···················· | ···················· | ···················· | ···················· |
| B | 572 hCoV-19/USA/WA-S237/2020\|EPI_ISL_430275\|2020-03-06 | ···················· | ···················· | ···················· | ···················· | ···················· | ···················· |
| B | 573 hCoV-19/USA/WA-S238/2020\|EPI_ISL_430276\|2020-03-02 | ···················· | ···················· | ···················· | ···················· | ···················· | ···················· |
| B | 574 hCoV-19/USA/WA-S239/2020\|EPI_ISL_430277\|2020-03-07 | ···················· | ···················· | ···················· | ···················· | ···················· | ···················· |
| B | 575 hCoV-19/USA/WA-S240/2020\|EPI_ISL_430278\|2020-03-06 | ···················· | ···················· | ···················· | ···················· | ···················· | ···················· |
| B | 576 hCoV-19/USA/WA-S241/2020\|EPI_ISL_430279\|2020-03-04 | ···················· | ···················· | ···················· | ···················· | ···················· | ···················· |
| B | 578 hCoV-19/USA/WA-S243/2020\|EPI_ISL_430281\|2020-02-29 | ···················· | ···················· | ···················· | ···················· | ···················· | ···················· |
| B | 579 hCoV-19/USA/WA-S244/2020\|EPI_ISL_430282\|2020-03-11 | ···················· | ···················· | ···················· | ···················· | ···················· | ···················· |
| B | 580 hCoV-19/USA/WA-S245/2020\|EPI_ISL_430283\|2020-03-02 | ···················· | ···················· | ···················· | ···················· | ···················· | ···················· |
| B | 581 hCoV-19/USA/WA-S246/2020\|EPI_ISL_430284\|2020-03-10 | ···················· | ···················· | ···················· | ···················· | ···················· | ···················· |
| B | 582 hCoV-19/USA/WA-S247/2020\|EPI_ISL_430285\|2020-03-01 | ···················· | ···················· | ···················· | ···················· | ···················· | ···················· |
| B | 583 hCoV-19/USA/WA-S248/2020\|EPI_ISL_430286\|2020-03-10 | ···················· | ···················· | ···················· | ···················· | ···················· | ···················· |
| B | 584 hCoV-19/USA/WA-S249/2020\|EPI_ISL_430287\|2020-03-01 | ···················· | ···················· | ···················· | ···················· | ···················· | ···················· |
| B | 585 hCoV-19/USA/WA-S250/2020\|EPI_ISL_430288\|2020-03-03 | ···················· | ···················· | ···················· | ···················· | ···················· | ···················· |
| B | 586 hCoV-19/USA/WA-S251/2020\|EPI_ISL_430289\|2020-03-11 | ···················· | ···················· | ···················· | ···················· | ···················· | ···················· |
| B | 587 hCoV-19/USA/WA-S252/2020\|EPI_ISL_430290\|2020-03-02 | ···················· | ···················· | ···················· | ···················· | ···················· | ···················· |
| B | 588 hCoV-19/USA/WA-S253/2020\|EPI_ISL_430291\|2020-03-07 | ···················· | ···················· | ···················· | ···················· | ···················· | ···················· |
| B | 589 hCoV-19/USA/WA-S254/2020\|EPI_ISL_430292\|2020-02-24 | ···················· | ···················· | ···················· | ···················· | ···················· | ···················· |
| B | 590 hCoV-19/USA/WA-S255/2020\|EPI_ISL_430293\|2020-03-08 | ···················· | ···················· | ···················· | ···················· | ···················· | ···················· |
| B | 591 hCoV-19/USA/WA-S256/2020\|EPI_ISL_430294\|2020-03-02 | ···················· | ···················· | ···················· | ···················· | ···················· | ···················· |
| B | 592 hCoV-19/USA/WA-S257/2020\|EPI_ISL_430295\|2020-02-25 | ···················· | ···················· | ···················· | ···················· | ···················· | ···················· |
| B | 593 hCoV-19/South Africa/R02827/2020\|EPI_ISL_430297\|20 | ···················· | ···················· | ···················· | ···················· | ···················· | ···················· |
| B | 594 hCoV-19/England/SHEF-C5D79/2020\|EPI_ISL_432812\|202 | ···················· | ···················· | ···················· | ···················· | ···················· | ···················· |
| B | 595 hCoV-19/England/SHEF-C5DD3/2020\|EPI_ISL_432813\|202 | ···················· | ·A·················· | ···················· | ···················· | ···················· | ···················· |
| B | 596 hCoV-19/Scotland/EDB909/2020\|EPI_ISL_432874\|2020-0 | ···················· | ···················· | ···················· | ···················· | ·············N······ | ···················· |
| B | 597 hCoV-19/Scotland/EDB910/2020\|EPI_ISL_432875\|2020-0 | ···················· | ···················· | ···················· | ···················· | ···················· | ···················· |
| B | 599 hCoV-19/Scotland/EDB914/2020\|EPI_ISL_432877\|2020-0 | ···················· | ···················· | ···················· | ···················· | ···················· | ···················· |
| B | 601 hCoV-19/Scotland/EDB921/2020\|EPI_ISL_432880\|2020-0 | ···················· | ···················· | ···················· | ···················· | ···················· | ···················· |
| B | 602 hCoV-19/Scotland/EDB922/2020\|EPI_ISL_432881\|2020-0 | ···················· | ···················· | ···················· | ···················· | ···················· | ···················· |
| B | 603 hCoV-19/Scotland/EDB924/2020\|EPI_ISL_432883\|2020-0 | ···················· | ···················· | ···················· | ···················· | ···················· | ···················· |
| B | 604 hCoV-19/Scotland/EDB926/2020\|EPI_ISL_432885\|2020-0 | ···················· | ·A·················· | ···················· | ···················· | ···················· | ···················· |
| B | 605 hCoV-19/Scotland/EDB927/2020\|EPI_ISL_432886\|2020-0 | ···················· | ···················· | ···················· | ···················· | ···················· | ···················· |
| B | 608 hCoV-19/Scotland/EDB945/2020\|EPI_ISL_432890\|2020-0 | ···················· | ···················· | ···················· | ···················· | ···················· | ···················· |
| B | 609 hCoV-19/Scotland/EDB964/2020\|EPI_ISL_432891\|2020-0 | ···················· | ·A·················· | ···················· | ···················· | ···N················ | ···················· |
| B | 610 hCoV-19/Scotland/EDB971/2020\|EPI_ISL_432892\|2020-0 | ···················· | ·A·················· | ···················· | ···················· | ···················· | ···················· |
| B | 611 hCoV-19/Scotland/EDB972/2020\|EPI_ISL_432893\|2020-0 | ···················· | ···················· | ···················· | ···················· | ···················· | ···················· |
| B | 612 hCoV-19/Scotland/EDB973/2020\|EPI_ISL_432894\|2020-0 | ···················· | ···················· | ···················· | ···················· | ···················· | ···················· |
| B | 613 hCoV-19/Scotland/EDB986/2020\|EPI_ISL_432895\|2020-0 | ···················· | ···················· | ···················· | ···················· | ···················· | ···················· |
| B | 615 hCoV-19/Scotland/EDB997/2020\|EPI_ISL_432898\|2020-0 | ···················· | ·A·················· | ···················· | ···················· | ···················· | ···················· |
| B | 616 hCoV-19/Scotland/EDB999/2020\|EPI_ISL_432899\|2020-0 | ···················· | ···················· | ···················· | ···················· | ···················· | ···················· |
| B | 617 hCoV-19/India/JK-NCDC-02326/2020\|EPI_ISL_435090\|20 | ···················· | ···················· | ···················· | ···················· | ···················· | ···················· |
| B | 618 hCoV-19/England/CAMB-7DFF2/2020\|EPI_ISL_442200\|202 | ···················· | ···················· | ···················· | ···················· | ···················· | ···················· |
| B | 619 hCoV-19/England/CAMB-7E0B2/2020\|EPI_ISL_442201\|202 | ···················· | ···················· | ···················· | ···················· | ···················· | ···················· |
| B | 620 hCoV-19/England/CAMB-7DE8C/2020\|EPI_ISL_442202\|202 | ···················· | ···················· | ···················· | ···················· | ···················· | ···················· |
| B | 621 hCoV-19/England/CAMB-7DFB6/2020\|EPI_ISL_442203\|202 | ···················· | ···················· | ···················· | ···················· | ···················· | ···················· |
| B | 622 hCoV-19/England/CAMB-7D62A/2020\|EPI_ISL_442204\|202 | ···················· | ···················· | ···················· | ···················· | ···················· | ···················· |
| B | 623 hCoV-19/England/CAMB-7D068/2020\|EPI_ISL_442206\|202 | ···················· | ···················· | ···················· | ···················· | ···················· | ···················· |
| B | 624 hCoV-19/England/CAMB-7BC1B/2020\|EPI_ISL_442207\|202 | ···················· | ···················· | ···················· | ···················· | ···················· | ···················· |
| B | 625 hCoV-19/England/CAMB-7CED8/2020\|EPI_ISL_442208\|202 | ···················· | ···················· | ···················· | ···················· | ···················· | ···················· |
| B | 626 hCoV-19/England/CAMB-7B6D1/2020\|EPI_ISL_442210\|202 | ···················· | ···················· | ···················· | ···················· | ···················· | ···················· |
| B | 627 hCoV-19/England/CAMB-7C755/2020\|EPI_ISL_442211\|202 | ···················· | ···················· | ···················· | ···················· | ···················· | ···················· |
| B | 628 hCoV-19/England/CAMB-7CB4A/2020\|EPI_ISL_442213\|202 | ···················· | ···················· | ···················· | ···················· | ···················· | ···················· |
| B | 629 hCoV-19/England/CAMB-7C737/2020\|EPI_ISL_442214\|202 | ···················· | ···················· | ···················· | ···················· | ···················· | ···················· |
| B | 630 hCoV-19/England/CAMB-7CD08/2020\|EPI_ISL_442215\|202 | ···················· | ···················· | ···················· | ···················· | ···················· | ···················· |
| B | 631 hCoV-19/England/CAMB-7D7BE/2020\|EPI_ISL_442216\|202 | ···················· | ···················· | ···················· | ···················· | ···················· | ···················· |
| B | 632 hCoV-19/England/CAMB-7D1CF/2020\|EPI_ISL_442217\|202 | ···················· | ···················· | ···················· | ···················· | ···················· | ···················· |
| B | 633 hCoV-19/England/CAMB-7CF7B/2020\|EPI_ISL_442224\|202 | ···················· | ···················· | ···················· | ···················· | ···················· | ···················· |
| B | 634 hCoV-19/England/CAMB-7DEAA/2020\|EPI_ISL_442225\|202 | ···················· | ···················· | ···················· | ···················· | ···················· | ···················· |
| B | 635 hCoV-19/England/CAMB-7DC0A/2020\|EPI_ISL_442226\|202 | ···················· | ···················· | ···················· | ···················· | ···················· | ···················· |
| B | 636 hCoV-19/England/CAMB-7CFB7/2020\|EPI_ISL_442229\|202 | ···················· | ···················· | ···················· | ···················· | ···················· | ···················· |
| B | 637 hCoV-19/England/CAMB-7CEC9/2020\|EPI_ISL_442230\|202 | ···················· | ···················· | ···················· | ···················· | ···················· | ···················· |
| B | 638 hCoV-19/England/CAMB-7D0C2/2020\|EPI_ISL_442231\|202 | ···················· | ···················· | ···················· | ···················· | ···················· | ···················· |
| B | 639 hCoV-19/England/CAMB-7CFC6/2020\|EPI_ISL_442232\|202 | ···················· | ···················· | ···················· | ···················· | ···················· | ···················· |
| B | 640 hCoV-19/England/CAMB-7D11A/2020\|EPI_ISL_442234\|202 | ···················· | ···················· | ···················· | ···················· | ···················· | ···················· |
| B | 641 hCoV-19/England/CAMB-7BAF3/2020\|EPI_ISL_442235\|202 | ···················· | A··················· | ···················· | ···················· | ···················· | ···················· |
| B | 642 hCoV-19/England/CAMB-7DF01/2020\|EPI_ISL_442236\|202 | ···················· | ···················· | ···················· | ···················· | ···················· | ···················· |

| | | | | | | | |
|---|---|---|---|---|---|---|---|
| B | 643 hCoV-19/England/CAMB-7C843/2020\|EPI_ISL_442237\|202 | .................... | .................... | ..................... | .................... | .................... | .................... |
| B | 644 hCoV-19/England/CAMB-7D3E7/2020\|EPI_ISL_442240\|202 | .................... | .................... | ..................... | .................... | .................... | .................... |
| B | 645 hCoV-19/England/CAMB-7D9B8/2020\|EPI_ISL_442241\|202 | .................... | .................... | ..................... | .................... | .................... | .................... |
| B | 646 hCoV-19/England/CAMB-7D578/2020\|EPI_ISL_442242\|202 | .................... | .................... | ..................... | .................... | .................... | .................... |
| B | 647 hCoV-19/England/CAMB-7C54C/2020\|EPI_ISL_442244\|202 | .................... | .................... | ..................... | .................... | .................... | .................... |
| B | 648 hCoV-19/England/CAMB-7C658/2020\|EPI_ISL_442246\|202 | .................... | .................... | ..................... | .................... | .................... | .................... |
| B | 649 hCoV-19/England/CAMB-7C825/2020\|EPI_ISL_442247\|202 | .................... | .................... | ..................... | .................... | .................... | .................... |
| B | 650 hCoV-19/England/CAMB-7C4F4/2020\|EPI_ISL_442248\|202 | .................... | .................... | ..................... | .................... | .................... | .................... |
| B | 651 hCoV-19/England/CAMB-7D7DC/2020\|EPI_ISL_442250\|202 | .................... | .................... | ..................... | .................... | .................... | .................... |
| B | 652 hCoV-19/England/CAMB-7CDCC/2020\|EPI_ISL_442251\|202 | .................... | .................... | ..................... | .................... | .................... | .................... |
| B | 653 hCoV-19/England/CAMB-7D38D/2020\|EPI_ISL_442252\|202 | .................... | .................... | ..................... | .................... | .................... | .................... |
| B | 654 hCoV-19/England/CAMB-7D39C/2020\|EPI_ISL_442253\|202 | .................... | .................... | ..................... | .................... | .................... | .................... |
| B | 655 hCoV-19/England/CAMB-7CE50/2020\|EPI_ISL_442254\|202 | .................... | .................... | ..................... | .................... | .................... | .................... |
| B | 656 hCoV-19/England/CAMB-7CD80/2020\|EPI_ISL_442255\|202 | .................... | .................... | ..................... | .................... | .................... | .................... |
| B | 657 hCoV-19/England/CAMB-7CF4E/2020\|EPI_ISL_442256\|202 | .................... | .................... | ..................... | .................... | .................... | .................... |
| B | 658 hCoV-19/England/CAMB-7CDBD/2020\|EPI_ISL_442257\|202 | .................... | .................... | ..................... | .................... | .................... | .................... |
| B | 659 hCoV-19/England/CAMB-7DCA0/2020\|EPI_ISL_442258\|202 | .................... | .................... | ..................... | .................... | .................... | .................... |
| B | 660 hCoV-19/England/CAMB-7B6FF/2020\|EPI_ISL_442259\|202 | .................... | .................... | ..................... | .................... | .................... | .................... |
| B | 661 hCoV-19/England/CAMB-7D350/2020\|EPI_ISL_442260\|202 | .................... | .................... | ..................... | .................... | .................... | .................... |
| B | 662 hCoV-19/England/CAMB-7D9D6/2020\|EPI_ISL_442261\|202 | .................... | .................... | ..................... | .................... | .................... | .................... |
| B | 663 hCoV-19/England/CAMB-7B729/2020\|EPI_ISL_442262\|202 | .................... | .................... | ..................... | .................... | .................... | .................... |
| B | 664 hCoV-19/England/CAMB-7D2F9/2020\|EPI_ISL_442263\|202 | .................... | .................... | ..................... | .................... | .................... | .................... |
| B | 665 hCoV-19/England/CAMB-7C60D/2020\|EPI_ISL_442265\|202 | .................... | .................... | ..................... | .................... | .................... | .................... |
| B | 666 hCoV-19/England/CAMB-7D4F3/2020\|EPI_ISL_442266\|202 | .................... | ....A............... | ..................... | .................... | .................... | .................... |
| B | 667 hCoV-19/England/CAMB-7BB2D/2020\|EPI_ISL_442268\|202 | .................... | .................... | ..................... | .................... | .................... | .................... |
| B | 668 hCoV-19/England/CAMB-7CFE4/2020\|EPI_ISL_442269\|202 | .................... | .................... | ..................... | .................... | .................... | .................... |
| B | 669 hCoV-19/England/CAMB-7CF8A/2020\|EPI_ISL_442270\|202 | .................... | .................... | ..................... | .................... | .................... | .................... |
| B | 670 hCoV-19/England/CAMB-7CF99/2020\|EPI_ISL_442272\|202 | .................... | .................... | ..................... | .................... | .................... | .................... |
| B | 671 hCoV-19/England/CAMB-7DCEC/2020\|EPI_ISL_442273\|202 | .................... | .................... | ..................... | .................... | .................... | .................... |
| B | 672 hCoV-19/England/CAMB-7D2DB/2020\|EPI_ISL_442274\|202 | .................... | .................... | ..................... | .................... | .................... | .................... |
| B | 673 hCoV-19/England/CAMB-7BAA8/2020\|EPI_ISL_442275\|202 | .................... | .................... | ..................... | .................... | .................... | .................... |
| B | 674 hCoV-19/England/CAMB-7D420/2020\|EPI_ISL_442276\|202 | .................... | .................... | ..................... | .................... | .................... | .................... |
| B | 675 hCoV-19/England/CAMB-7C430/2020\|EPI_ISL_442277\|202 | .................... | .................... | ..................... | .................... | .................... | .................... |
| B | 676 hCoV-19/England/CAMB-7C47C/2020\|EPI_ISL_442278\|202 | .................... | .................... | ..................... | .................... | .................... | .................... |
| B | 677 hCoV-19/England/CAMB-8307E/2020\|EPI_ISL_443522\|202 | .................... | .................... | ..................... | .................... | .................... | .................... |
| B | 678 hCoV-19/England/CAMB-836A9/2020\|EPI_ISL_443523\|202 | .................... | .................... | ..................... | .................... | .................... | .................... |
| B | 679 hCoV-19/England/CAMB-826F5/2020\|EPI_ISL_443524\|202 | .................... | .................... | ..................... | .................... | .................... | .................... |
| B | 680 hCoV-19/England/CAMB-8458D/2020\|EPI_ISL_443525\|202 | .................... | .................... | ..................... | .................... | .................... | .................... |
| B | 681 hCoV-19/England/CAMB-82394/2020\|EPI_ISL_443526\|202 | .................... | .................... | ..................... | .................... | .................... | .................... |
| B | 682 hCoV-19/England/CAMB-83418/2020\|EPI_ISL_443527\|202 | .................... | .................... | ..................... | .................... | .................... | .................... |
| B | 683 hCoV-19/England/CAMB-8280E/2020\|EPI_ISL_443528\|202 | .................... | .................... | ..................... | .................... | .................... | .................... |
| B | 684 hCoV-19/England/CAMB-1AA40B/2020\|EPI_ISL_443529\|20 | .................... | .................... | ..................... | .................... | .................... | .................... |
| B | 685 hCoV-19/England/CAMB-8291A/2020\|EPI_ISL_443530\|202 | .................... | .................... | ..................... | .................... | .................... | .................... |
| B | 686 hCoV-19/England/CAMB-82640/2020\|EPI_ISL_443531\|202 | .................... | .................... | ..................... | .................... | .................... | .................... |
| B | 687 hCoV-19/England/CAMB-83612/2020\|EPI_ISL_443532\|202 | .................... | .................... | ..................... | .................... | .................... | .................... |
| B | 688 hCoV-19/England/CAMB-82437/2020\|EPI_ISL_443533\|202 | .................... | .................... | ..................... | .................... | .................... | .................... |
| B | 689 hCoV-19/England/CAMB-844CC/2020\|EPI_ISL_443534\|202 | .................... | .................... | ..................... | .................... | .................... | .................... |
| B | 690 hCoV-19/England/CAMB-82974/2020\|EPI_ISL_443535\|202 | .................... | .................... | ..................... | .................... | .................... | .................... |
| B | 691 hCoV-19/England/CAMB-82DD2/2020\|EPI_ISL_443536\|202 | .................... | .................... | ..................... | .................... | .................... | .................... |
| B | 692 hCoV-19/England/CAMB-843A1/2020\|EPI_ISL_443537\|202 | .................... | .................... | ..................... | .................... | .................... | .................... |
| B | 693 hCoV-19/England/CAMB-1AA3A4/2020\|EPI_ISL_443538\|20 | .................... | .................... | ..................... | .................... | .................... | .................... |
| B | 694 hCoV-19/England/CAMB-822B5/2020\|EPI_ISL_443540\|202 | .................... | .................... | ..................... | .................... | .................... | .................... |
| B | 695 hCoV-19/England/CAMB-82FEB/2020\|EPI_ISL_443545\|202 | .................... | .................... | ..................... | .................... | .................... | .................... |
| B | 696 hCoV-19/England/CAMB-1AA32C/2020\|EPI_ISL_443546\|20 | .................... | .................... | ..................... | .................... | .................... | .................... |
| B | 697 hCoV-19/England/CAMB-83111/2020\|EPI_ISL_443548\|202 | .................... | .................... | ..................... | .................... | .................... | .................... |
| B | 698 hCoV-19/England/CAMB-838A3/2020\|EPI_ISL_443549\|202 | .................... | .................... | ..................... | .................... | .................... | .................... |
| B | 699 hCoV-19/England/CAMB-833C0/2020\|EPI_ISL_443557\|202 | .................... | .................... | ..................... | .................... | .................... | .................... |
| B | 700 hCoV-19/England/CAMB-825AD/2020\|EPI_ISL_443558\|202 | .................... | .................... | ..................... | .................... | .................... | .................... |
| B | 701 hCoV-19/England/CAMB-823A3/2020\|EPI_ISL_443559\|202 | .................... | .................... | ..................... | .................... | .................... | .................... |
| B | 702 hCoV-19/England/CAMB-82DC3/2020\|EPI_ISL_443560\|202 | .................... | .................... | ..................... | .................... | .................... | .................... |
| B | 703 hCoV-19/England/CAMB-8313F/2020\|EPI_ISL_443561\|202 | .................... | .................... | ..................... | .................... | .................... | .................... |
| B | 704 hCoV-19/England/CAMB-82570/2020\|EPI_ISL_443562\|202 | .................... | .................... | ..................... | .................... | .................... | .................... |
| B | 705 hCoV-19/England/CAMB-8424A/2020\|EPI_ISL_443564\|202 | .................... | .................... | ..................... | .................... | .................... | .................... |
| B | 706 hCoV-19/England/CAMB-843FC/2020\|EPI_ISL_443566\|202 | .................... | .................... | ..................... | .................... | .................... | .................... |
| B | 707 hCoV-19/England/CAMB-82DF0/2020\|EPI_ISL_443567\|202 | .................... | .................... | ..................... | .................... | .................... | .................... |
| B | 708 hCoV-19/England/CAMB-8382B/2020\|EPI_ISL_443568\|202 | .................... | .................... | ..................... | .................... | .................... | .................... |
| B | 709 hCoV-19/England/CAMB-84523/2020\|EPI_ISL_443569\|202 | .................... | .................... | ..................... | .................... | .................... | .................... |
| B | 710 hCoV-19/England/CAMB-83436/2020\|EPI_ISL_443571\|202 | .................... | .................... | ..................... | .................... | .................... | .................... |
| B | 711 hCoV-19/England/CAMB-837B5/2020\|EPI_ISL_443572\|202 | .................... | .................... | ..................... | .................... | .................... | .................... |
| B | 712 hCoV-19/England/CAMB-843B0/2020\|EPI_ISL_443574\|202 | .................... | .................... | ..................... | .................... | .................... | .................... |
| B | 713 hCoV-19/England/CAMB-8318A/2020\|EPI_ISL_443575\|202 | .................... | .................... | ..................... | .................... | .................... | .................... |
| B | 714 hCoV-19/England/CAMB-82859/2020\|EPI_ISL_443576\|202 | .................... | .................... | ..................... | .................... | .................... | .................... |
| B | 715 hCoV-19/England/CAMB-82956/2020\|EPI_ISL_443577\|202 | .................... | .................... | ..................... | .................... | .................... | .................... |
| B | 716 hCoV-19/England/CAMB-82D87/2020\|EPI_ISL_443578\|202 | .................... | .................... | ..................... | .................... | .................... | .................... |
| B | 717 hCoV-19/England/CAMB-826E6/2020\|EPI_ISL_443579\|202 | .................... | .................... | ..................... | .................... | .................... | .................... |
| B | 718 hCoV-19/England/CAMB-827D4/2020\|EPI_ISL_443581\|202 | .................... | .................... | ..................... | .................... | .................... | .................... |
| B | 719 hCoV-19/England/CAMB-8317B/2020\|EPI_ISL_443582\|202 | .................... | .................... | ..................... | .................... | .................... | .................... |
| B | 720 hCoV-19/England/CAMB-8266E/2020\|EPI_ISL_443583\|202 | .................... | .................... | ..................... | .................... | .................... | .................... |

| | | | | | | | | | |
|---|---|---|---|---|---|---|---|---|---|
| B | 721 | hCoV-19/England/CAMB-823C1/2020\|EPI_ISL_443585\|202 | ···················· | ···················· | ···················· | ···················· | ···················· | ···················· | |
| B | 722 | hCoV-19/England/CAMB-82701/2020\|EPI_ISL_443586\|202 | ···················· | ···················· | ···················· | ···················· | ···················· | ···················· | |
| B | 723 | hCoV-19/England/CAMB-82297/2020\|EPI_ISL_443587\|202 | ···················· | ···················· | ···················· | ···················· | ···················· | ···················· | |
| B | 724 | hCoV-19/England/CAMB-8276B/2020\|EPI_ISL_443588\|202 | ···················· | ···················· | ···················· | ···················· | ···················· | ···················· | |
| B | 725 | hCoV-19/England/CAMB-844BD/2020\|EPI_ISL_443589\|202 | ···················· | ···················· | ···················· | ···················· | ···················· | ···················· | |
| B | 726 | hCoV-19/England/CAMB-824CE/2020\|EPI_ISL_443590\|202 | ···················· | ···················A | ···················· | ···················· | ···················· | ···················· | 1 |
| B | 727 | hCoV-19/England/CAMB-82543/2020\|EPI_ISL_443591\|202 | ···················· | ···················· | ···················· | ···················· | ···················· | ···················· | |
| B | 728 | hCoV-19/England/CAMB-82446/2020\|EPI_ISL_443592\|202 | ···················· | ···················· | ···················· | ···················· | ···················· | ···················· | |
| B | 729 | hCoV-19/England/CAMB-83A34/2020\|EPI_ISL_443593\|202 | ···················· | ···················· | ···················· | ···················· | ···················· | ···················· | |
| B | 730 | hCoV-19/England/CAMB-837C4/2020\|EPI_ISL_443594\|202 | ···················· | ···················· | ···················· | ···················· | ···················· | ···················· | |
| B | 731 | hCoV-19/England/CAMB-83348/2020\|EPI_ISL_443595\|202 | ···················· | ···················· | ···················· | ···················· | ···················· | ···················· | |
| B | 732 | hCoV-19/England/CAMB-83885/2020\|EPI_ISL_443598\|202 | ···················· | ···················· | ···················· | ···················· | ···················· | ···················· | |
| B | 733 | hCoV-19/England/CAMB-838C1/2020\|EPI_ISL_443599\|202 | ···················· | ···················· | ···················· | ···················· | ···················· | ···················· | |
| B | 734 | hCoV-19/Denmark/ALAB-SSI-1387/2020\|EPI_ISL_444853\| | ···················· | ···················· | ···················· | ···················· | ···················· | ···················· | |
| B | 735 | hCoV-19/Denmark/ALAB-SSI-1388/2020\|EPI_ISL_444854\| | ···················· | ···················· | ···················· | ···················· | ···················· | ···················· | |
| B | 736 | hCoV-19/Denmark/ALAB-SSI-1390/2020\|EPI_ISL_444855\| | ···················· | ···················· | ···················· | ···················· | ···················· | ···················· | |
| B | 737 | hCoV-19/Denmark/ALAB-SSI-1391/2020\|EPI_ISL_444856\| | ···················· | ···················· | ···················· | ···················· | ···················· | ···················· | |
| B | 738 | hCoV-19/Denmark/ALAB-SSI-1393/2020\|EPI_ISL_444857\| | ···················· | ···················· | ·············N······ | ···················· | ···················· | ···················· | |
| B | 739 | hCoV-19/Denmark/ALAB-SSI-681/2020\|EPI_ISL_444859\|2 | ···················· | ···················· | ···················· | ···················· | ···················· | ···················· | |
| B | 740 | hCoV-19/Denmark/ALAB-SSI-682/2020\|EPI_ISL_444860\|2 | ···················· | ···················· | ···················· | ···················· | ···················· | ···················· | |
| B | 741 | hCoV-19/Denmark/ALAB-SSI-684/2020\|EPI_ISL_444861\|2 | ···················· | ···················· | ···················· | ···················· | ···················· | ···················· | |
| B | 742 | hCoV-19/Denmark/ALAB-SSI-687/2020\|EPI_ISL_444862\|2 | ···················· | ···················· | ···················· | ···················· | ···················· | ···················· | |
| B | 743 | hCoV-19/Denmark/ALAB-SSI-694/2020\|EPI_ISL_444863\|2 | ···················· | ···················· | ···················· | ···················· | ···················· | ···················· | |
| B | 744 | hCoV-19/Denmark/ALAB-SSI-700/2020\|EPI_ISL_444864\|2 | ···················· | ···················· | ···················· | ···················· | ···················· | ···················· | |
| B | 745 | hCoV-19/Denmark/ALAB-SSI-705/2020\|EPI_ISL_444866\|2 | ···················· | ···················· | ···················· | ···················· | ···················· | ···················· | |
| B | 746 | hCoV-19/Denmark/ALAB-SSI-707/2020\|EPI_ISL_444867\|2 | ···················· | ···················· | ···················· | ···················· | ···················· | ···················· | |
| B | 747 | hCoV-19/Denmark/ALAB-SSI-711/2020\|EPI_ISL_444869\|2 | ···················· | ···················· | ···················· | ···················· | ···················· | ···················· | |
| B | 748 | hCoV-19/Denmark/ALAB-SSI-713/2020\|EPI_ISL_444870\|2 | ···················· | ···················· | ···················· | ···················· | ···················· | ···················· | |
| B | 749 | hCoV-19/Denmark/ALAB-SSI-718/2020\|EPI_ISL_444872\|2 | ···················· | ···················· | ···················· | ···················· | ···················· | ···················· | |
| B | 750 | hCoV-19/Denmark/ALAB-SSI-719/2020\|EPI_ISL_444873\|2 | ···················· | ···················· | ···················· | ···················· | ···················· | ···················· | |
| B | 751 | hCoV-19/Denmark/ALAB-SSI-723/2020\|EPI_ISL_444874\|2 | ···················· | ···················· | ···················· | ···················· | ···················· | ···················· | |
| B | 752 | hCoV-19/Denmark/ALAB-SSI-724/2020\|EPI_ISL_444875\|2 | ···················· | ···················· | ···················· | ···················· | ···················· | ···················· | |
| B | 753 | hCoV-19/Denmark/ALAB-SSI-726/2020\|EPI_ISL_444877\|2 | ···················· | ···················· | ···················· | ···················· | ···················· | ···················· | |
| B | 754 | hCoV-19/Denmark/ALAB-SSI-728/2020\|EPI_ISL_444879\|2 | ···················· | ···················· | ···················· | ···················· | ···················· | ···················· | |
| B | 755 | hCoV-19/Denmark/ALAB-SSI-747/2020\|EPI_ISL_444887\|2 | ···················· | ···················· | ···················· | ···················· | ···················· | ···················· | |
| B | 756 | hCoV-19/Denmark/ALAB-SSI-751/2020\|EPI_ISL_444888\|2 | ···················· | ···················· | ···················· | ···················· | ···················· | ···················· | |
| B | 757 | hCoV-19/Denmark/ALAB-SSI-756/2020\|EPI_ISL_444890\|2 | ···················· | ···················· | ···················· | ···················· | ···················· | ···················· | |
| B | 758 | hCoV-19/Denmark/ALAB-SSI-757/2020\|EPI_ISL_444891\|2 | ···················· | ···················· | ···················· | ···················· | ···················· | ···················· | |
| B | 759 | hCoV-19/Denmark/ALAB-SSI-761/2020\|EPI_ISL_444892\|2 | ···················· | ···················· | ···················· | ···················· | ···················· | ···················· | |
| B | 760 | hCoV-19/Denmark/ALAB-SSI-762/2020\|EPI_ISL_444893\|2 | ···················· | ···················· | ···················· | ···················· | ···················· | ···················· | |
| B | 761 | hCoV-19/Denmark/ALAB-SSI-765/2020\|EPI_ISL_444894\|2 | ···················· | ···················· | ···················· | ···················· | ···················· | ···················· | |
| B | 762 | hCoV-19/Denmark/ALAB-SSI-767/2020\|EPI_ISL_444895\|2 | ···················· | ···················· | ···················· | ···················· | ···················· | ···················· | |
| B | 763 | hCoV-19/Denmark/ALAB-SSI-769/2020\|EPI_ISL_444896\|2 | ···················· | ···················· | ···················· | ···················· | ···················· | ···················· | |
| B | 764 | hCoV-19/Denmark/ALAB-SSI-770/2020\|EPI_ISL_444897\|2 | ···················· | ···················· | ···················· | ···················· | ···················· | ···················· | |
| B | 765 | hCoV-19/Denmark/ALAB-SSI-773/2020\|EPI_ISL_444898\|2 | ···················· | ···················· | ···················· | ···················· | ···················· | ···················· | |
| B | 766 | hCoV-19/Denmark/ALAB-SSI-774/2020\|EPI_ISL_444899\|2 | ···················· | ···················· | ···················· | ···················· | ···················· | ···················· | |
| B | 767 | hCoV-19/USA/MI-MDHHS-SC20273/2020\|EPI_ISL_447071\|2 | ···················· | ···················· | ···················· | ···················· | ···················· | ···················· | |
| B | 768 | hCoV-19/USA/MI-MDHHS-SC20279/2020\|EPI_ISL_447077\|2 | ···················· | ···················· | ···················· | ···················· | ···················· | ···················· | |
| B | 769 | hCoV-19/USA/WA-S166/2020\|EPI_ISL_430204\|2020-03-18 | ···················· | ···················· | ···················· | ···················· | ···················· | ···················· | |
| B | 770 | hCoV-19/USA/WA-S167/2020\|EPI_ISL_430205\|2020-03-18 | ···················· | ···················· | ···················· | ···················· | ···················· | ···················· | |
| B | 771 | hCoV-19/USA/WA-S168/2020\|EPI_ISL_430206\|2020-03-18 | ···················· | ···················· | ···················· | ···················· | ···················· | ···················· | |
| B | 772 | hCoV-19/USA/WA-S169/2020\|EPI_ISL_430207\|2020-03-18 | ···················· | ···················· | ···················· | ···················· | ···················· | ···················· | |
| B | 773 | hCoV-19/USA/WA-S170/2020\|EPI_ISL_430208\|2020-03-17 | ···················· | ···················· | ···················· | ···················· | ···················· | ···················· | |
| B | 774 | hCoV-19/USA/WA-S171/2020\|EPI_ISL_430209\|2020-03-19 | ···················· | ···················· | ···················· | ···················· | ···················· | ···················· | |
| B | 775 | hCoV-19/USA/WA-S172/2020\|EPI_ISL_430210\|2020-03-19 | ···················· | ···················· | ···················· | ···················· | ···················· | ···················· | |
| B | 776 | hCoV-19/USA/WA-S173/2020\|EPI_ISL_430211\|2020-03-19 | ···················· | ···················· | ···················· | ···················· | ···················· | ···················· | |
| B | 777 | hCoV-19/England/SHEF-C5359/2020\|EPI_ISL_432802\|202 | ···················· | ···················· | ···················· | ···················· | ···················· | ···················· | |
| B | 778 | hCoV-19/England/SHEF-C56AB/2020\|EPI_ISL_432804\|202 | ···················· | ···················· | ···················· | ···················· | ···················· | ···················· | |
| B | 779 | hCoV-19/England/SHEF-C6789/2020\|EPI_ISL_432825\|202 | ···················· | ···················· | ···················· | ···················· | ···················· | ···················· | |
| B | 780 | hCoV-19/England/SHEF-C6C11/2020\|EPI_ISL_432828\|202 | ···················· | ···················· | ···················· | ···················· | ···················· | ···················· | |
| B | 781 | hCoV-19/England/SHEF-C6EA2/2020\|EPI_ISL_432831\|202 | ···················· | ···················· | ···················· | ···················· | ···················· | ···················· | |
| B | 782 | hCoV-19/England/SHEF-C715D/2020\|EPI_ISL_432836\|202 | ···················· | ···················· | ···················· | ···················· | ···················· | ···················· | |
| B | 783 | hCoV-19/England/SHEF-C7296/2020\|EPI_ISL_432837\|202 | ···················· | ···················· | ···················· | ···················· | ···················· | ···················· | |
| B | 784 | hCoV-19/England/SHEF-C7937/2020\|EPI_ISL_432850\|202 | ···················· | ···················· | ···················· | ···················· | ···················· | ···················· | |
| B | 785 | hCoV-19/England/SHEF-C81D4/2020\|EPI_ISL_432859\|202 | ···················· | ···················· | ···················· | ···················· | ···················· | ···················· | |
| B | 786 | hCoV-19/England/SHEF-C8338/2020\|EPI_ISL_432861\|202 | ···················· | ····A··············· | ···················· | ···················· | ···················· | ···················· | |
| B | 787 | hCoV-19/England/SHEF-C83ED/2020\|EPI_ISL_432864\|202 | ···················· | ···················· | ···················· | ···················· | ···················· | ···················· | |
| B | 788 | hCoV-19/England/SHEF-C8532/2020\|EPI_ISL_432867\|202 | ···················· | ···················· | ···················· | ···················· | ···················· | ···················· | |
| B | 789 | hCoV-19/Scotland/EDB894/2020\|EPI_ISL_432868\|2020-0 | ···················· | ·A·················· | ···················· | ···················· | ···················· | ···················· | |
| B | 790 | hCoV-19/Scotland/EDB895/2020\|EPI_ISL_432869\|2020-0 | ···················· | ···················· | ···················· | ···················· | ···················· | ···················· | |
| B | 791 | hCoV-19/Scotland/EDB896/2020\|EPI_ISL_432870\|2020-0 | ···················· | ···················· | ···················· | ···················· | ···················· | ···················· | |
| B | 792 | hCoV-19/Scotland/EDB897/2020\|EPI_ISL_432871\|2020-0 | ···················· | ···················· | ···················· | ···················· | ···················· | ···················· | |
| B | 793 | hCoV-19/Scotland/EDB899/2020\|EPI_ISL_432872\|2020-0 | ···················· | ···················· | ···················· | ···················· | ···················· | ···················· | |
| B | 794 | hCoV-19/Scotland/EDB901/2020\|EPI_ISL_432873\|2020-0 | ···················· | ·A·················· | ···················· | ···················· | ···················· | ···················· | |
| B | 795 | hCoV-19/USA/TX-HMH0416/2020\|EPI_ISL_435000\|2020-04 | ···················· | ···················· | ···················· | ···················· | ···················· | ···················· | |
| B | 796 | hCoV-19/USA/TX-HMH0417/2020\|EPI_ISL_435001\|2020-04 | ···················· | ···················· | ···················· | ···················· | ···················· | ···················· | |
| B | 797 | hCoV-19/USA/TX-HMH0418/2020\|EPI_ISL_435002\|2020-04 | ···················· | ···················· | ···················· | ···················· | ···················· | ···················· | |
| B | 798 | hCoV-19/USA/TX-HMH0420/2020\|EPI_ISL_435003\|2020-04 | ···················· | ···················· | ···················· | ···················· | ···················· | ···················· | |

| | | | | | | | |
|---|---|---|---|---|---|---|---|
| B | 799 hCoV-19/USA/TX-HMH0422/2020\|EPI_ISL_435005\|2020-04 | ···················· | ···················· | ···················· | ···················· | ···················· | ···················· |
| B | 800 hCoV-19/USA/TX-HMH0423/2020\|EPI_ISL_435006\|2020-04 | ···················· | ···················· | ···················· | ···················· | ···················· | ···················· |
| B | 801 hCoV-19/USA/TX-HMH0424/2020\|EPI_ISL_435007\|2020-04 | ···················· | ···················· | ···················· | ···················· | ···················· | ···················· |
| B | 802 hCoV-19/USA/TX-HMH0425/2020\|EPI_ISL_435008\|2020-04 | ···················· | ···················· | ···················· | ···················· | ···················· | ···················· |
| B | 803 hCoV-19/USA/TX-HMH0426/2020\|EPI_ISL_435009\|2020-04 | ···················· | ···················· | ···················· | ···················· | ···················· | ···················· |
| B | 804 hCoV-19/USA/TX-HMH0427/2020\|EPI_ISL_435010\|2020-04 | ···················· | ···················· | ···················· | ···················· | ···················· | ···················· |
| B | 805 hCoV-19/USA/TX-HMH0428/2020\|EPI_ISL_435011\|2020-04 | ···················· | ···················· | ···················· | ···················· | ···················· | ···················· |
| B | 806 hCoV-19/USA/TX-HMH0429/2020\|EPI_ISL_435012\|2020-04 | ···················· | ···················· | ···················· | ···················· | ···················· | ···················· |
| B | 807 hCoV-19/USA/TX-HMH0430/2020\|EPI_ISL_435013\|2020-04 | ···················· | ···················· | ···················· | ···················· | ···················· | ···················· |
| B | 808 hCoV-19/USA/TX-HMH0431/2020\|EPI_ISL_435014\|2020-04 | ···················· | ···················· | ···················· | ···················· | ···················· | ···················· |
| B | 809 hCoV-19/USA/TX-HMH0432/2020\|EPI_ISL_435015\|2020-04 | ···················· | ···················· | ···················· | ···················· | ···················· | ···················· |
| B | 810 hCoV-19/USA/TX-HMH0433/2020\|EPI_ISL_435016\|2020-04 | ···················· | ···················· | ···················· | ···················· | ···················· | ···················· |
| B | 811 hCoV-19/USA/TX-HMH0434/2020\|EPI_ISL_435017\|2020-04 | ···················· | ···················· | ···················· | ···················· | ···················· | ···················· |
| B | 812 hCoV-19/USA/TX-HMH0435/2020\|EPI_ISL_435018\|2020-04 | ···················· | ···················· | ···················· | ···················· | ···················· | ···················· |
| B | 813 hCoV-19/USA/TX-HMH0436/2020\|EPI_ISL_435019\|2020-04 | ···················· | ···················· | ···················· | ···················· | ···················· | ···················· |
| B | 814 hCoV-19/USA/TX-HMH0437/2020\|EPI_ISL_435020\|2020-04 | ···················· | ···················· | ···················· | ···················· | ···················· | ···················· |
| B | 815 hCoV-19/USA/TX-HMH0438/2020\|EPI_ISL_435021\|2020-04 | ···················· | ···················· | ···················· | ···················· | ···················· | ···················· |
| B | 816 hCoV-19/USA/TX-HMH0439/2020\|EPI_ISL_435022\|2020-04 | ···················· | ···················· | ···················· | ···················· | ···················· | ···················· |
| B | 817 hCoV-19/USA/TX-HMH0440/2020\|EPI_ISL_435023\|2020-04 | ···················· | ···················· | ···················· | ···················· | ···················· | ···················· |
| B | 818 hCoV-19/USA/TX-HMH0441/2020\|EPI_ISL_435024\|2020-04 | ···················· | ···················· | ···················· | ···················· | ···················· | ···················· |
| B | 819 hCoV-19/USA/TX-HMH0449/2020\|EPI_ISL_435028\|2020-04 | ···················· | ···················· | ···················· | ···················· | ···················· | ···················· |
| B | 820 hCoV-19/USA/TX-HMH0450/2020\|EPI_ISL_435029\|2020-04 | ···················· | ···················· | ···················· | ···················· | ···················· | ···················· |
| B | 821 hCoV-19/India/GJ-GBRC3/2020\|EPI_ISL_435050\|2020-04 | ···················· | ···················· | ···················· | ···················· | ···················· | ···················· |
| B | 822 hCoV-19/India/GJ-GBRC4/2020\|EPI_ISL_435051\|2020-04 | ···················· | ···················· | ···················· | ···················· | ···················· | ···················· |
| B | 823 hCoV-19/India/GJ-GBRC5/2020\|EPI_ISL_435052\|2020-04 | ···················· | ···················· | ···················· | ···················· | ···················· | ···················· |
| B | 824 hCoV-19/India/GJ-GBRC6/2020\|EPI_ISL_435053\|2020-04 | ···················· | ···················· | ···················· | ···················· | ···················· | ···················· |
| B | 825 hCoV-19/India/GJ-GBRC7/2020\|EPI_ISL_435054\|2020-04 | ···················· | ···················· | ···················· | ···················· | ···················· | ···················· |
| B | 826 hCoV-19/India/GJ-GBRC8/2020\|EPI_ISL_435055\|2020-04 | ···················· | ·A·················· | ···················· | ···················· | ···················· | ···A················ |
| B | 827 hCoV-19/India/GJ-GBRC9/2020\|EPI_ISL_435056\|2020-04 | ···················· | ···················· | ···················· | ···················· | ···················· | ···················· |
| B | 828 hCoV-19/India/UP-NCDC-01257/2020\|EPI_ISL_435060\|20 | ···················· | ···················· | ···················· | ···················· | ···················· | ···················· |
| B | 829 hCoV-19/India/DL-NCDC-01537/2020\|EPI_ISL_435061\|20 | ···················· | ···················· | ···················· | ···················· | ···················· | ···················· |
| B | 830 hCoV-19/India/PB-NCDC-01538/2020\|EPI_ISL_435062\|20 | ···················· | ·N·················· | ···················· | ···················· | ···················· | ···················· |
| B | 831 hCoV-19/India/DL-NCDC-01326/2020\|EPI_ISL_435063\|20 | ···················· | ···················· | ···················· | ···················· | ···················· | ···················· |
| B | 832 hCoV-19/India/DL-NCDC-01705/2020\|EPI_ISL_435064\|20 | ···················· | ···················· | ···················· | ···················· | ···················· | ···················· |
| B | 833 hCoV-19/India/DL-NCDC-01475/2020\|EPI_ISL_435065\|20 | ···················· | ·A·················· | ···················· | ···················· | ···················· | ···A················ |
| B | 834 hCoV-19/India/DL-NCDC-01711/2020\|EPI_ISL_435066\|20 | ···················· | ·A·················· | ···················· | ···················· | ···················· | ···A················ |
| B | 835 hCoV-19/India/DL-NCDC-01712/2020\|EPI_ISL_435067\|20 | ···················· | ·A·················· | ···················· | ···················· | ···················· | ···A················ |
| B | 836 hCoV-19/India/DL-NCDC-01757/2020\|EPI_ISL_435068\|20 | ···················· | ·A·················· | ···················· | ···················· | ···················· | ···A················ |
| B | 837 hCoV-19/India/DL-NCDC-01744/2020\|EPI_ISL_435069\|20 | ···················· | ·A·················· | ···················· | ···················· | ···················· | ···A················ |
| B | 838 hCoV-19/India/DL-NCDC-01710/2020\|EPI_ISL_435070\|20 | ···················· | ···················· | ···················· | ···················· | ···················· | ···················· |
| B | 839 hCoV-19/India/DL-NCDC-02105/2020\|EPI_ISL_435071\|20 | ···················· | ···················· | ···················· | ···················· | ···················· | ···················· |
| B | 840 hCoV-19/India/DL-NCDC-02155/2020\|EPI_ISL_435072\|20 | ···················· | ···················· | ···················· | ···················· | ··········N········· | ···················· |
| B | 841 hCoV-19/India/WB-NCDC-02250/2020\|EPI_ISL_435074\|20 | ···················· | ···················· | ···················· | ···················· | ···················· | ···················· |
| B | 842 hCoV-19/India/TN-NCDC-02311/2020\|EPI_ISL_435078\|20 | ···················· | ···················· | ···················· | ···················· | ·············N······ | ···················· |
| B | 843 hCoV-19/India/TN-NCDC-02334/2020\|EPI_ISL_435080\|20 | ···················· | ···················· | ···················· | ···················· | ···················· | ···················· |
| B | 844 hCoV-19/India/WB-NCDC-02248/2020\|EPI_ISL_435081\|20 | ···················· | ···················· | ···················· | ···················· | ···················· | ···················· |
| B | 845 hCoV-19/India/UP-NCDC-02322/2020\|EPI_ISL_435082\|20 | ···················· | ···················· | ···················· | ···················· | ···················· | ···················· |
| B | 846 hCoV-19/India/TN-NCDC-02242/2020\|EPI_ISL_435083\|20 | ···················· | ···················· | ···················· | ···················· | ···················· | ···················· |
| B | 847 hCoV-19/India/TN-NCDC-02309/2020\|EPI_ISL_435084\|20 | ···················· | ···················· | ···················· | ···················· | ···················· | ···················· |
| B | 848 hCoV-19/India/MH-NCDC-02310/2020\|EPI_ISL_435085\|20 | ···················· | ···················· | ···················· | ···················· | ···················· | ···················· |
| B | 849 hCoV-19/India/MH-NCDC-02315/2020\|EPI_ISL_435086\|20 | ···················· | ···················· | ···················· | ···················· | ···················· | ···················· |
| B | 850 hCoV-19/India/TN-NCDC-02333/2020\|EPI_ISL_435087\|20 | ···················· | ···················· | ···················· | ···················· | ···················· | ···················· |
| B | 851 hCoV-19/India/OR-NCDC-02336/2020\|EPI_ISL_435088\|20 | ···················· | ···················· | ···················· | ···················· | ···················· | ···················· |
| B | 852 hCoV-19/India/TN-NCDC-02245/2020\|EPI_ISL_435091\|20 | ···················· | ···················· | ···················· | ···················· | ···················· | ···················· |
| B | 853 hCoV-19/India/TN-NCDC-02244/2020\|EPI_ISL_435092\|20 | ···················· | ···················· | ···················· | ···················· | ·······N···W········ | ···················· |
| B | 854 hCoV-19/India/TN-NCDC-02312/2020\|EPI_ISL_435093\|20 | ···················· | ···················· | ···················· | ···················· | ··········N········· | ···················· |
| B | 855 hCoV-19/India/TN-NCDC-02327/2020\|EPI_ISL_435094\|20 | ···················· | ···················· | ···················· | ···················· | ···················· | ···················· |
| B | 856 hCoV-19/India/TN-NCDC-02328/2020\|EPI_ISL_435095\|20 | ···················· | ···················· | ···················· | ···················· | ···················· | ···················· |
| B | 857 hCoV-19/India/TN-NCDC-02332/2020\|EPI_ISL_435096\|20 | ···················· | ···················· | ···················· | ···················· | ···················· | ···················· |
| B | 858 hCoV-19/India/WB-NCDC-02252/2020\|EPI_ISL_435097\|20 | ···················· | ···················· | ···················· | ···················· | ···················· | ···················· |
| B | 859 hCoV-19/India/UN-NCDC-02240/2020\|EPI_ISL_435098\|20 | ···················· | ···················· | ···················· | ···················· | ···················· | ···················· |
| B | 860 hCoV-19/India/UP-NCDC-02320/2020\|EPI_ISL_435099\|20 | ···················· | ···················· | ···················· | ···················· | ··········N········· | ···················· |
| B | 861 hCoV-19/Spain/VC-FISABIO-222/2020\|EPI_ISL_436317\|2 | ···················· | ···················· | ···················· | ···················· | ···················· | ···················· |
| B | 862 hCoV-19/Spain/VC-FISABIO-223/2020\|EPI_ISL_436318\|2 | ···················· | ···················· | ···················· | ···················· | ···················· | ···················· |
| B | 864 hCoV-19/Spain/VC-FISABIO-231/2020\|EPI_ISL_436326\|2 | ···················· | ···················· | ···················· | ···················· | ···················· | ···················· |
| B | 865 hCoV-19/Spain/VC-FISABIO-232/2020\|EPI_ISL_436327\|2 | ···················· | ···················· | ···················· | ···················· | ···················· | ···················· |
| B | 866 hCoV-19/Spain/VC-FISABIO-233/2020\|EPI_ISL_436328\|2 | ···················· | ···················· | ···················· | ···················· | ···················· | ···················· |
| B | 867 hCoV-19/Spain/VC-FISABIO-235/2020\|EPI_ISL_436330\|2 | ···················· | ···················· | ···················· | ···················· | ···················· | ···················· |
| B | 868 hCoV-19/Spain/VC-FISABIO-240/2020\|EPI_ISL_436335\|2 | ···················· | ···················· | ···················· | ···················· | ···················· | ···················· |
| B | 870 hCoV-19/Spain/VC-FISABIO-242/2020\|EPI_ISL_436337\|2 | ···················· | ···················· | ···················· | ···················· | ···················· | ···················· |
| B | 872 hCoV-19/Spain/VC-FISABIO-247/2020\|EPI_ISL_436342\|2 | ···················· | ···················· | ···················· | ···················· | ···················· | ···················· |
| B | 873 hCoV-19/Spain/VC-FISABIO-250/2020\|EPI_ISL_436345\|2 | ···················· | ···················· | ···················· | ···················· | ···················· | ···················· |
| B | 874 hCoV-19/Spain/VC-FISABIO-251/2020\|EPI_ISL_436346\|2 | ···················· | ···················· | ···················· | ···················· | ···················· | ···················· |
| B | 877 hCoV-19/Spain/VC-FISABIO-261/2020\|EPI_ISL_436356\|2 | ···················· | ···················· | ···················· | ···················· | ···················· | ···················· |
| B | 878 hCoV-19/Spain/VC-FISABIO-285/2020\|EPI_ISL_436380\|2 | ···················· | ···················· | ···················· | ···················· | ···················· | ···················· |
| B | 879 hCoV-19/Spain/VC-FISABIO-287/2020\|EPI_ISL_436382\|2 | ···················· | ···················· | ···················· | ···················· | ···················· | ···················· |
| B | 880 hCoV-19/Spain/VC-FISABIO-288/2020\|EPI_ISL_436383\|2 | ···················· | ···················· | ···················· | ···················· | ···················· | ···················· |
| B | 881 hCoV-19/Spain/VC-FISABIO-289/2020\|EPI_ISL_436384\|2 | ···················· | ···················· | ···················· | ···················· | ···················· | ···················· |

| | | | | | | | | |
|---|---|---|---|---|---|---|---|---|
| B | 882 hCoV-19/Spain/VC-FISABIO-291/2020\|EPI_ISL_436386\|2 | ···················· | ···················· | ····················· | ···················· | ···················· | ···················· | |
| B | 883 hCoV-19/Spain/VC-FISABIO-292/2020\|EPI_ISL_436387\|2 | ···················· | ···················· | ····················· | ···················· | ···················· | ···················· | |
| B | 884 hCoV-19/Spain/VC-FISABIO-293/2020\|EPI_ISL_436388\|2 | ···················· | ···················· | ····················· | ···················· | ···················· | ···················· | |
| B | 885 hCoV-19/Spain/VC-FISABIO-294/2020\|EPI_ISL_436389\|2 | ···················· | ···················· | ····················· | ···················· | ···················· | ···················· | |
| B | 886 hCoV-19/Spain/VC-FISABIO-295/2020\|EPI_ISL_436390\|2 | ···················· | ···················· | ····················· | ···················· | ···················· | ···················· | |
| B | 887 hCoV-19/Spain/VC-FISABIO-296/2020\|EPI_ISL_436391\|2 | ···················· | ···················· | ····················· | ···················· | ···················· | ···················· | |
| B | 888 hCoV-19/Spain/VC-FISABIO-302/2020\|EPI_ISL_436397\|2 | ···················· | ···················· | ····················· | ···················· | ···················· | ···················· | |
| B | 889 hCoV-19/Denmark/ALAB-SSI-136/2020\|EPI_ISL_437648\|2 | ···················· | ···················· | ····················· | ···················· | ···················· | ···················· | |
| B | 890 hCoV-19/Denmark/ALAB-SSI-137/2020\|EPI_ISL_437649\|2 | ···················· | ···················· | ····················· | ···················· | ···················· | ···················· | |
| B | 891 hCoV-19/Denmark/ALAB-SSI-138/2020\|EPI_ISL_437650\|2 | ···················· | ···················· | ····················· | ···················· | ···················· | ···················· | |
| B | 892 hCoV-19/Denmark/ALAB-SSI-1406/2020\|EPI_ISL_437651\| | ···················· | ···················· | ····················· | ···················· | ···················· | ···················· | |
| B | 893 hCoV-19/Denmark/ALAB-SSI-1421/2020\|EPI_ISL_437653\| | ···················· | ···················· | ····················· | ···················· | ···················· | ···················· | |
| B | 894 hCoV-19/Denmark/ALAB-SSI-1427/2020\|EPI_ISL_437654\| | ···················· | ···················· | ····················· | ···················· | ···················· | ···················· | |
| B | 895 hCoV-19/Denmark/ALAB-SSI-1434/2020\|EPI_ISL_437655\| | ···················· | ···················· | ····················· | ···················· | ···················· | ···················· | |
| B | 896 hCoV-19/Denmark/ALAB-SSI-1498/2020\|EPI_ISL_437656\| | ···················· | ···················· | ····················· | ···················· | ···················· | ···················· | |
| B | 897 hCoV-19/Denmark/ALAB-SSI-242/2020\|EPI_ISL_437657\|2 | ···················· | ···················· | ····················· | ···················· | ···················· | ···················· | |
| B | 898 hCoV-19/Denmark/ALAB-SSI-243/2020\|EPI_ISL_437658\|2 | ···················· | ···················· | ····················· | ···················· | ···················· | ···················· | |
| B | 899 hCoV-19/Denmark/ALAB-SSI-244/2020\|EPI_ISL_437659\|2 | ···················· | ···················· | ····················· | ···················· | ···················· | ···················· | |
| B | 900 hCoV-19/Denmark/ALAB-SSI-245/2020\|EPI_ISL_437660\|2 | ···················· | ···················· | ····················· | ···················· | ···················· | ···················· | |
| B | 901 hCoV-19/Denmark/ALAB-SSI-246/2020\|EPI_ISL_437661\|2 | ···················· | ···················· | ····················· | ···················· | ···················· | ···················· | |
| B | 902 hCoV-19/Denmark/ALAB-SSI-248/2020\|EPI_ISL_437663\|2 | ···················· | ···················· | ····················· | ···················· | ···················· | ···················· | |
| B | 903 hCoV-19/Denmark/ALAB-SSI-251/2020\|EPI_ISL_437664\|2 | ···················· | ···················· | ····················· | ···················· | ···················· | ···················· | |
| B | 904 hCoV-19/Denmark/ALAB-SSI-745/2020\|EPI_ISL_437667\|2 | ···················· | ···················· | ····················· | ···················· | ···················· | ···················· | |
| B | 905 hCoV-19/Denmark/ALAB-SSI-748/2020\|EPI_ISL_437668\|2 | ···················· | ···················· | ····················· | ···················· | ···················· | ···················· | |
| B | 906 hCoV-19/Denmark/ALAB-SSI-783/2020\|EPI_ISL_437669\|2 | ···················· | ···················· | ····················· | ···················· | ···················· | ···················· | |
| B | 907 hCoV-19/Denmark/ALAB-SSI-786/2020\|EPI_ISL_437670\|2 | ···················· | ···················· | ····················· | ···················· | ···················· | ···················· | |
| B | 908 hCoV-19/Denmark/ALAB-SSI-800/2020\|EPI_ISL_437672\|2 | ···················· | ···················· | ····················· | ···················· | ···················· | ···················· | |
| B | 909 hCoV-19/Denmark/ALAB-SSI-817/2020\|EPI_ISL_437674\|2 | ···················· | ···················· | ····················· | ···················· | ···················· | ···················· | |
| B | 910 hCoV-19/Denmark/ALAB-SSI-825/2020\|EPI_ISL_437676\|2 | ···················· | ···················· | ····················· | ···················· | ···················· | ···················· | |
| B | 911 hCoV-19/Denmark/ALAB-SSI-834/2020\|EPI_ISL_437677\|2 | ···················· | ···················· | ····················· | ···················· | ···················· | ···················· | |
| B | 912 hCoV-19/Denmark/ALAB-SSI-845/2020\|EPI_ISL_437678\|2 | ···················· | ···················· | ····················· | ···················· | ···················· | ···················· | |
| B | 913 hCoV-19/Denmark/ALAB-SSI-864/2020\|EPI_ISL_437679\|2 | ···················· | ···················· | ····················· | ···················· | ···················· | ···················· | |
| B | 914 hCoV-19/Denmark/ALAB-SSI-991/2020\|EPI_ISL_437683\|2 | ···················· | ···················· | ····················· | ···················· | ···················· | ···················· | |
| B | 915 hCoV-19/Ireland/un-NVRL-20W266/2020\|EPI_ISL_437684 | ···················· | ···················· | ····················· | ···················· | ···················· | ···················· | |
| B | 916 hCoV-19/Ireland/un-NVRL-20W267/2020\|EPI_ISL_437685 | ···················· | ···················· | ····················· | ···················· | ···················· | ···················· | |
| B | 917 hCoV-19/Ireland/un-NVRL-20W268/2020\|EPI_ISL_437686 | ···················· | ···················· | ····················· | ···················· | ···················· | ···················· | |
| B | 918 hCoV-19/Ireland/un-NVRL-20W269/2020\|EPI_ISL_437687 | ···················· | ···················· | ····················· | ···················· | ···················· | ···················· | |
| B | 919 hCoV-19/Ireland/un-NVRL-20W270/2020\|EPI_ISL_437688 | ···················· | ···················· | ····················· | ···················· | ···················· | ···················· | |
| B | 920 hCoV-19/France/IDF-CIBU200107/2020\|EPI_ISL_437689\| | ···················· | ···················· | ····················· | ···················· | ···················· | ···················· | |
| B | 921 hCoV-19/Scotland/CVR2789/2020\|EPI_ISL_438979\|2020- | ···················· | ···················A | ····················· | ···················· | ···················· | ···················· | 1 |
| B | 922 hCoV-19/Scotland/CVR2802/2020\|EPI_ISL_438980\|2020- | ···················· | ···················· | ····················· | ···················· | ···················· | ···················· | |
| B | 923 hCoV-19/Scotland/CVR2803/2020\|EPI_ISL_438981\|2020- | ···················· | ···················· | ····················· | ···················· | ···················· | ···················· | |
| B | 924 hCoV-19/Scotland/CVR2805/2020\|EPI_ISL_438982\|2020- | ···················· | ···················· | ····················· | ···················· | ···················· | ···················· | |
| B | 925 hCoV-19/Scotland/CVR2810/2020\|EPI_ISL_438983\|2020- | ···················· | ···················· | ····················· | ···················· | ···················· | ···················· | |
| B | 926 hCoV-19/Scotland/CVR2812/2020\|EPI_ISL_438984\|2020- | ···················· | ···················· | ····················· | ···················· | ···················· | ···················· | |
| B | 927 hCoV-19/Scotland/CVR2814/2020\|EPI_ISL_438985\|2020- | ···················· | ···················· | ····················· | ···················· | ···················· | ···················· | |
| B | 928 hCoV-19/Scotland/CVR2819/2020\|EPI_ISL_438986\|2020- | ···················· | ···················· | ····················· | ···················· | ···················· | ···················· | |
| B | 929 hCoV-19/Scotland/CVR2906/2020\|EPI_ISL_438995\|2020- | ···················· | ···················· | ····················· | ···················· | ···················· | ···················· | |
| B | 930 hCoV-19/Scotland/CVR2921/2020\|EPI_ISL_438998\|2020- | ···················· | ···················· | ····················· | ···················· | ···················· | ···················· | |
| B | 931 hCoV-19/Scotland/CVR2937/2020\|EPI_ISL_438999\|2020- | ···················· | ···················· | ···········T·T······· | ···················· | ···················· | ···················· | |
| B | 932 hCoV-19/England/CAMB-834AF/2020\|EPI_ISL_443500\|202 | ···················· | ···················· | ····················· | ···················· | ···················· | ···················· | |
| B | 933 hCoV-19/England/CAMB-82561/2020\|EPI_ISL_443501\|202 | ···················· | ···················· | ····················· | ···················· | ···················· | ···················· | |
| B | 934 hCoV-19/England/CAMB-1AA4B0/2020\|EPI_ISL_443502\|20 | ···················· | ···················· | ····················· | ···················· | ···················· | ···················· | |
| B | 935 hCoV-19/England/CAMB-8226A/2020\|EPI_ISL_443503\|202 | ···················· | ···················· | ····················· | ···················· | ···················· | ···················· | |
| B | 936 hCoV-19/England/CAMB-8267D/2020\|EPI_ISL_443504\|202 | ···················· | ···················· | ····················· | ···················· | ···················· | ···················· | |
| B | 937 hCoV-19/England/CAMB-84514/2020\|EPI_ISL_443505\|202 | ···················· | ···················· | ····················· | ···················· | ···················· | ···················· | |
| B | 938 hCoV-19/England/CAMB-826D7/2020\|EPI_ISL_443507\|202 | ···················· | ···················· | ····················· | ···················· | ···················· | ···················· | |
| B | 939 hCoV-19/England/CAMB-831F3/2020\|EPI_ISL_443509\|202 | ···················· | ···················· | ····················· | ···················· | ···················· | ···················· | |
| B | 940 hCoV-19/England/CAMB-84453/2020\|EPI_ISL_443510\|202 | ···················· | ···················· | ····················· | ···················· | ···················· | ···················· | |
| B | 941 hCoV-19/England/CAMB-84541/2020\|EPI_ISL_443511\|202 | ···················· | ···················· | ····················· | ···················· | ···················· | ···················· | |
| B | 942 hCoV-19/England/CAMB-83409/2020\|EPI_ISL_443512\|202 | ···················· | ···················· | ····················· | ···················· | ···················· | ···················· | |
| B | 943 hCoV-19/England/CAMB-1AA4DE/2020\|EPI_ISL_443514\|20 | ···················· | ···················· | ····················· | ···················· | ···················· | ···················· | |
| B | 944 hCoV-19/England/CAMB-1AA3C2/2020\|EPI_ISL_443515\|20 | ···················· | ···················· | ····················· | ···················· | ···················· | ···················· | |
| B | 945 hCoV-19/England/CAMB-1AA359/2020\|EPI_ISL_443517\|20 | ···················· | ···················· | ····················· | ···················· | ···················· | ···················· | |
| B | 946 hCoV-19/England/CAMB-845E7/2020\|EPI_ISL_443519\|202 | ···················· | ···················· | ····················· | ···················· | ···················· | ···················· | |
| B | 947 hCoV-19/Denmark/ALAB-SSI-1235/2020\|EPI_ISL_444817\| | ···················· | ···················· | ····················· | ···················· | ···················· | ···················· | |
| B | 948 hCoV-19/Denmark/ALAB-SSI-1238/2020\|EPI_ISL_444818\| | ···················· | ···················· | ····················· | ···················· | ···················· | ···················· | |
| B | 949 hCoV-19/Denmark/ALAB-SSI-1239/2020\|EPI_ISL_444819\| | ···················· | ···················· | ····················· | ···················· | ···················· | ···················· | |
| B | 950 hCoV-19/Denmark/ALAB-SSI-1241/2020\|EPI_ISL_444820\| | ···················· | ···················· | ····················· | ···················· | ···················· | ···················· | |
| B | 951 hCoV-19/Denmark/ALAB-SSI-1243/2020\|EPI_ISL_444822\| | ···················· | ···················· | ····················· | ···················· | ···················· | ···················· | |
| B | 952 hCoV-19/Denmark/ALAB-SSI-1244/2020\|EPI_ISL_444823\| | ···················· | ···················· | ····················· | ···················· | ···················· | ···················· | |
| B | 953 hCoV-19/Denmark/ALAB-SSI-1245/2020\|EPI_ISL_444824\| | ···················· | ···················· | ····················· | ···················· | ···················· | ···················· | |
| B | 954 hCoV-19/Denmark/ALAB-SSI-1246/2020\|EPI_ISL_444825\| | ···················· | ···················· | ····················· | ···················· | ···················· | ···················· | |
| B | 955 hCoV-19/Denmark/ALAB-SSI-1248/2020\|EPI_ISL_444826\| | ···················· | ···················· | ····················· | ···················· | ···················· | ···················· | |
| B | 956 hCoV-19/Denmark/ALAB-SSI-1249/2020\|EPI_ISL_444827\| | ···················· | ···················· | ····················· | ···················· | ···················· | ···················· | |
| B | 957 hCoV-19/Denmark/ALAB-SSI-1250/2020\|EPI_ISL_444828\| | ···················· | ···················· | ····················· | ···················· | ···················· | ···················· | |
| B | 958 hCoV-19/Denmark/ALAB-SSI-1252/2020\|EPI_ISL_444829\| | ···················· | ···················· | ····················· | ···················· | ···················· | ···················· | |
| B | 959 hCoV-19/Denmark/ALAB-SSI-1257/2020\|EPI_ISL_444831\| | ···················· | ···················· | ····················· | ···················· | ···················· | ···················· | |

| | | | | | | | | |
|---|---|---|---|---|---|---|---|---|
| B | 960 | hCoV-19/Denmark/ALAB-SSI-1260/2020\|EPI_ISL_444832\| | ···················· | ···················· | ···················· | ···················· | ···················· | ···················· |
| B | 961 | hCoV-19/Denmark/ALAB-SSI-1261/2020\|EPI_ISL_444833\| | ···················· | ···················· | ···················· | ···················· | ···················· | ···················· |
| B | 962 | hCoV-19/Denmark/ALAB-SSI-1262/2020\|EPI_ISL_444834\| | ···················· | ···················· | ···················· | ···················· | ···················· | ···················· |
| B | 963 | hCoV-19/Denmark/ALAB-SSI-1263/2020\|EPI_ISL_444835\| | ···················· | ···················· | ···················· | ···················· | ···················· | ···················· |
| B | 964 | hCoV-19/Denmark/ALAB-SSI-1264/2020\|EPI_ISL_444836\| | ···················· | ···················· | ···················· | ···················· | ···················· | ···················· |
| B | 965 | hCoV-19/Denmark/ALAB-SSI-1265/2020\|EPI_ISL_444837\| | ···················· | ···················· | ···················· | ···················· | ···················· | ···················· |
| B | 966 | hCoV-19/Denmark/ALAB-SSI-1268/2020\|EPI_ISL_444838\| | ···················· | ···················· | ···················· | ···················· | ···················· | ···················· |
| B | 967 | hCoV-19/Denmark/ALAB-SSI-1269/2020\|EPI_ISL_444839\| | ···················· | ···················· | ···················· | ···················· | ···················· | ···················· |
| B | 968 | hCoV-19/Denmark/ALAB-SSI-1270/2020\|EPI_ISL_444840\| | ···················· | ···················· | ···················· | ···················· | ···················· | ···················· |
| B | 969 | hCoV-19/Denmark/ALAB-SSI-1272/2020\|EPI_ISL_444841\| | ···················· | ···················· | ···················· | ···················· | ···················· | ···················· |
| B | 970 | hCoV-19/Denmark/ALAB-SSI-1282/2020\|EPI_ISL_444844\| | ···················· | ···················· | ···················· | ···················· | ···················· | ···················· |
| B | 971 | hCoV-19/Denmark/ALAB-SSI-1288/2020\|EPI_ISL_444846\| | ···················· | ···················· | ···················· | ···················· | ···················· | ···················· |
| B | 972 | hCoV-19/Denmark/ALAB-SSI-1292/2020\|EPI_ISL_444848\| | ···················· | ···················· | ···················· | ···················· | ···················· | ···················· |
| B | 973 | hCoV-19/Denmark/ALAB-SSI-1378/2020\|EPI_ISL_444849\| | ···················· | ···················· | ···················· | ···················· | ···················· | ···················· |
| B | 974 | hCoV-19/Denmark/ALAB-SSI-1382/2020\|EPI_ISL_444850\| | ···················· | ···················· | ···················· | ···················· | ···················· | ···················· |
| B | 975 | hCoV-19/Denmark/ALAB-SSI-1386/2020\|EPI_ISL_444852\| | ···················· | ···················· | ···················· | ···················· | ···················· | ···················· |
| B | 977 | hCoV-19/India/GJ-GBRC56a/2020\|EPI_ISL_447030\|2020- | ···················· | ·········A·········· | ···················· | ···················· | ···················· | ···················· |
| B | 978 | hCoV-19/India/GJ-GBRC56b/2020\|EPI_ISL_447031\|2020- | ···················· | ···················· | ···················· | ···················· | ···················· | ···················· |
| B | 979 | hCoV-19/India/GJ-GBRC57/2020\|EPI_ISL_447032\|2020-0 | ···················· | ···················· | ···················· | ···················· | ···················· | ···················· |
| B | 980 | hCoV-19/India/GJ-GBRC58/2020\|EPI_ISL_447033\|2020-0 | ···················· | ·A·················· | ···················· | ···················· | ···················· | ··A················· |
| B | 981 | hCoV-19/India/GJ-GBRC59/2020\|EPI_ISL_447034\|2020-0 | ···················· | ·A·················· | ···················· | ···················· | ···················· | ··A················· |
| B | 982 | hCoV-19/India/GJ-GBRC60/2020\|EPI_ISL_447035\|2020-0 | ···················· | ·A·················· | ···················· | ···················· | ···················· | ··A················· |
| B | 983 | hCoV-19/India/GJ-GBRC61/2020\|EPI_ISL_447036\|2020-0 | ···················· | ···················· | ···················· | ···················· | ···················· | ···················· |
| B | 984 | hCoV-19/India/GJ-GBRC62/2020\|EPI_ISL_447037\|2020-0 | ···················· | ···················· | ···················· | ···················· | ···················· | ···················· |
| B | 985 | hCoV-19/India/GJ-GBRC63/2020\|EPI_ISL_447038\|2020-0 | ···················· | ···················· | ···················· | ···················· | ···················· | ···················· |
| B | 986 | hCoV-19/India/GJ-GBRC64/2020\|EPI_ISL_447039\|2020-0 | ···················· | ·A·················· | ···················· | ···················· | ···················· | ··A················· |
| B | 987 | hCoV-19/India/GJ-GBRC65a/2020\|EPI_ISL_447040\|2020- | ···················· | ·A·················· | ···················· | ···················· | ···················· | ··A················· |
| B | 988 | hCoV-19/India/GJ-GBRC65b/2020\|EPI_ISL_447041\|2020- | ···················· | ···················· | ···················· | ···················· | ···················· | ··A················· |
| B | 989 | hCoV-19/India/GJ-GBRC66/2020\|EPI_ISL_447042\|2020-0 | ···················· | ···················· | ···················· | ···················· | ···················· | ···················· |
| B | 990 | hCoV-19/India/GJ-GBRC67a/2020\|EPI_ISL_447043\|2020- | ···················· | ·A·················· | ···················· | ···················· | ···················· | ··A················· |
| B | 991 | hCoV-19/India/GJ-GBRC67b/2020\|EPI_ISL_447044\|2020- | ···················· | ·A·················· | ···················· | ···················· | ···················· | ··A················· |
| B | 992 | hCoV-19/India/GJ-GBRC68/2020\|EPI_ISL_447045\|2020-0 | ···················· | ·A·················· | ···················· | ···················· | ···················· | ··A················· |
| B | 993 | hCoV-19/India/GJ-GBRC69/2020\|EPI_ISL_447046\|2020-0 | ···················· | ·A·················· | ···················· | ···················· | ···················· | ··A················· |
| B | 994 | hCoV-19/India/GJ-GBRC70/2020\|EPI_ISL_447047\|2020-0 | ···················· | ···················· | ···················· | ···················· | ···················· | ···················· |
| B | 995 | hCoV-19/India/GJ-GBRC71/2020\|EPI_ISL_447048\|2020-0 | ···················· | ·A·················· | ···················· | ···················· | ···················· | ··A················· |
| B | 996 | hCoV-19/India/GJ-GBRC72a/2020\|EPI_ISL_447049\|2020- | ···················· | ·A·················· | ···················· | ···················· | ···················· | ··A················· |
| B | 997 | hCoV-19/India/GJ-GBRC72b/2020\|EPI_ISL_447050\|2020- | ···················· | ·A·················· | ···················· | ···················· | ···················· | ··A················· |
| B | 998 | hCoV-19/India/GJ-GBRC73/2020\|EPI_ISL_447051\|2020-0 | ···················· | ·A·················· | ···················· | ···················· | ···················· | ··A················· |
| B | 999 | hCoV-19/India/GJ-GBRC74/2020\|EPI_ISL_447052\|2020-0 | ···················· | ···················· | ···················· | ···················· | ···················· | ···················· |
| B | 1000 | hCoV-19/India/GJ-GBRC75/2020\|EPI_ISL_447053\|2020-0 | ···················· | ···················· | ···················· | ···················· | ···················· | ···················· |
| B | 1001 | hCoV-19/Romania/279068/2020\|EPI_ISL_447054\|2020-04 | ···················· | ···················· | ···················· | ···················· | ···················· | ···················· |
| B | 1002 | hCoV-19/Georgia/Tb-6598/2020\|EPI_ISL_447055\|2020-0 | ···················· | ···················· | ···················· | ···················· | ···················· | ···················· |
| B | 1003 | hCoV-19/Georgia/Tb-6572/2020\|EPI_ISL_447056\|2020-0 | ···················· | ···················· | ···················· | ···················· | ···················· | ···················· |
| B | 1004 | hCoV-19/USA/MI-MDHHS-SC20192/2020\|EPI_ISL_447057\|2 | ···················· | ···················· | ···················· | ···················· | ···················· | ···················· |
| B | 1005 | hCoV-19/USA/MI-MDHHS-SC20260/2020\|EPI_ISL_447059\|2 | ···················· | ···················· | ···················· | ···················· | ···················· | ···················· |
| B | 1006 | hCoV-19/USA/MI-MDHHS-SC20264/2020\|EPI_ISL_447063\|2 | ···················· | ···················· | ···················· | ···················· | ···················· | ···················· |
| B | 1007 | hCoV-19/USA/MI-MDHHS-SC20266/2020\|EPI_ISL_447064\|2 | ···················· | ···················· | ···················· | ···················· | ···················· | ···················· |
| B | 1008 | hCoV-19/USA/MI-MDHHS-SC20270/2020\|EPI_ISL_447068\|2 | ···················· | ···················· | ···················· | ···················· | ···················· | ···················· |
| B | 1009 | hCoV-19/USA/MI-MDHHS-SC20271/2020\|EPI_ISL_447069\|2 | ···················· | ···················· | ···················· | ···················· | ···················· | ···················· |
| B | 1010 | hCoV-19/USA/MI-MDHHS-SC20274/2020\|EPI_ISL_447072\|2 | ···················· | ···················· | ···················· | ···················· | ···················· | ···················· |
| B | 1011 | hCoV-19/USA/MI-MDHHS-SC20283/2020\|EPI_ISL_447081\|2 | ···················· | ···················· | ···················· | ···················· | ···················· | ···················· |
| B | 1012 | hCoV-19/USA/MI-MDHHS-SC20284/2020\|EPI_ISL_447082\|2 | ···················· | ···················· | ···················· | ···················· | ···················· | ···················· |
| B | 1013 | hCoV-19/USA/MI-MDHHS-SC20285/2020\|EPI_ISL_447083\|2 | ···················· | ···················· | ···················· | ···················· | ···················· | ···················· |
| B | 1014 | hCoV-19/USA/MI-MDHHS-SC20291/2020\|EPI_ISL_447088\|2 | ···················· | ···················· | ···················· | ···················· | ···················· | ···················· |
| B | 1015 | hCoV-19/USA/MI-MDHHS-SC20292/2020\|EPI_ISL_447089\|2 | ···················· | ···················· | ···················· | ···················· | ···················· | ···················· |
| B | 1016 | hCoV-19/USA/MI-MDHHS-SC20293/2020\|EPI_ISL_447090\|2 | ···················· | ···················· | ···················· | ···················· | ···················· | ···················· |
| B | 1017 | hCoV-19/USA/MI-MDHHS-SC20294/2020\|EPI_ISL_447091\|2 | ···················· | ···················· | ···················· | ···················· | ···················· | ···················· |
| B | 1018 | hCoV-19/USA/MI-MDHHS-SC20295/2020\|EPI_ISL_447092\|2 | ···················· | ···················· | ···················· | ···················· | ···················· | ···················· |
| B | 1019 | hCoV-19/USA/MI-MDHHS-SC20296/2020\|EPI_ISL_447093\|2 | ···················· | ···················· | ···················· | ···················· | ···················· | ···················· |
| B | 1020 | hCoV-19/USA/MI-MDHHS-SC20297/2020\|EPI_ISL_447094\|2 | ···················· | ···················· | ···················· | ···················· | ···················· | ···················· |
| B | 1021 | hCoV-19/USA/MI-MDHHS-SC20298/2020\|EPI_ISL_447095\|2 | ···················· | ···················· | ···················· | ···················· | ···················· | ···················· |
| B | 1022 | hCoV-19/USA/MI-MDHHS-SC20299/2020\|EPI_ISL_447096\|2 | ···················· | ···················· | ···················· | ···················· | ···················· | ···················· |
| B | 1023 | hCoV-19/USA/MI-MDHHS-SC20301/2020\|EPI_ISL_447098\|2 | ···················· | ···················· | ···················· | ···················· | ···················· | ···················· |
| B | 1024 | hCoV-19/USA/MI-MDHHS-SC20302/2020\|EPI_ISL_447099\|2 | ···················· | ···················· | ···················· | ···················· | ···················· | ···················· |
| B | 1025 | hCoV-19/USA/WA-S142/2020\|EPI_ISL_430179\|2020-03-13 | ···················· | ···················· | ···················· | ···················· | ···················· | ···················· |
| B | 1026 | hCoV-19/USA/WA-S143/2020\|EPI_ISL_430180\|2020-03-15 | ···················· | ···················· | ···················· | ···················· | ···················· | ···················· |
| B | 1027 | hCoV-19/USA/WA-S144/2020\|EPI_ISL_430181\|2020-03-15 | ···················· | ···················· | ···················· | ···················· | ···················· | ···················· |
| B | 1028 | hCoV-19/USA/WA-S145/2020\|EPI_ISL_430182\|2020-03-15 | ···················· | ···················· | ···················· | ···················· | ···················· | ···················· |
| B | 1029 | hCoV-19/USA/WA-S146/2020\|EPI_ISL_430183\|2020-03-14 | ···················· | ···················· | ···················· | ···················· | ···················· | ···················· |
| B | 1030 | hCoV-19/USA/WA-S147/2020\|EPI_ISL_430184\|2020-03-14 | ···················· | ···················· | ···················· | ···················· | ···················· | ···················· |
| B | 1031 | hCoV-19/USA/WA-S148/2020\|EPI_ISL_430185\|2020-03-13 | ···················· | ···················· | ···················· | ···················· | ···················· | ···················· |
| B | 1032 | hCoV-19/USA/WA-S149/2020\|EPI_ISL_430186\|2020-03-14 | ···················· | ···················· | ···················· | ···················· | ···················· | ···················· |
| B | 1033 | hCoV-19/USA/WA-S150/2020\|EPI_ISL_430187\|2020-03-14 | ···················· | ···················· | ···················· | ···················· | ···················· | ···················· |
| B | 1034 | hCoV-19/USA/WA-S151/2020\|EPI_ISL_430188\|2020-03-16 | ···················· | ···················· | ···················· | ···················· | ···················· | ···················· |
| B | 1035 | hCoV-19/USA/WA-S152/2020\|EPI_ISL_430189\|2020-03-16 | ···················· | ···················· | ···················· | ···················· | ···················· | ···················· |
| B | 1036 | hCoV-19/USA/WA-S153/2020\|EPI_ISL_430190\|2020-03-16 | ···················· | ···················· | ···················· | ···················· | ···················· | ···················· |
| B | 1037 | hCoV-19/USA/WA-S154/2020\|EPI_ISL_430191\|2020-03-16 | ···················· | ···················· | ···················· | ···················· | ···················· | ···················· |
| B | 1038 | hCoV-19/USA/WA-S155/2020\|EPI_ISL_430192\|2020-03-16 | ···················· | ···················· | ···················· | ···················· | ···················· | ···················· |

| | | | | | | | | | |
|---|---|---|---|---|---|---|---|---|---|
| B | 1039 | hCoV-19/USA/WA-S156/2020\|EPI_ISL_430193\|2020-03-16 | ···················· | ···················· | ···················· | ···················· | ···················· | ···················· | |
| B | 1040 | hCoV-19/USA/WA-S158/2020\|EPI_ISL_430196\|2020-03-16 | ···················· | ···················· | ···················· | ···················· | ···················· | ···················· | |
| B | 1041 | hCoV-19/USA/WA-S159/2020\|EPI_ISL_430197\|2020-03-16 | ···················· | ···················· | ···················· | ···················· | ···················· | ···················· | |
| B | 1042 | hCoV-19/USA/WA-S160/2020\|EPI_ISL_430198\|2020-03-17 | ···················· | ···················· | ···················· | ···················· | ···················· | ···················· | |
| B | 1043 | hCoV-19/USA/WA-S161/2020\|EPI_ISL_430199\|2020-03-17 | ···················· | ···················· | ···················· | ···················· | ···················· | ···················· | |
| B | 1044 | hCoV-19/England/SHEF-CCC93/2020\|EPI_ISL_432533\|202 | ···················· | ···················· | ···················· | ···················· | ···················· | ···················· | |
| B | 1045 | hCoV-19/England/SHEF-CD05A/2020\|EPI_ISL_432539\|202 | ···················· | ·A·················· | ···················· | ···················· | ···················· | ···················· | |
| B | 1046 | hCoV-19/England/SHEF-CD2AF/2020\|EPI_ISL_432541\|202 | ···················· | ···················· | ···················· | ···················· | ···················· | ···················· | |
| B | 1047 | hCoV-19/England/SHEF-CD315/2020\|EPI_ISL_432542\|202 | ···················· | ···················· | ···················· | ···················· | ···················· | ···················· | |
| B | 1048 | hCoV-19/England/SHEF-CD52E/2020\|EPI_ISL_432547\|202 | ···················· | ···················· | ···················· | ···················· | ···················· | ···················· | |
| B | 1049 | hCoV-19/England/SHEF-CD737/2020\|EPI_ISL_432548\|202 | ···················· | ···················· | ···················· | ···················· | ···················· | ···················· | |
| B | 1050 | hCoV-19/England/SHEF-CD89E/2020\|EPI_ISL_432549\|202 | ···················· | ···················· | ···················· | ···················· | ···················· | ···················· | |
| B | 1051 | hCoV-19/England/SHEF-CD97D/2020\|EPI_ISL_432551\|202 | ···················· | ···················· | ···················· | ···················· | ···················· | ···················· | |
| B | 1052 | hCoV-19/England/CAMB-7ACC1/2020\|EPI_ISL_433864\|202 | ···················· | ···················· | ···················· | ···················· | ···················· | ···················· | |
| B | 1053 | hCoV-19/England/CAMB-7ACD0/2020\|EPI_ISL_433865\|202 | ···················· | ···················· | ···················· | ···················· | ·····T·············· | ···················· | |
| B | 1054 | hCoV-19/England/CAMB-7ACFE/2020\|EPI_ISL_433866\|202 | ···················· | ···················A | ···················· | ···················· | ···················· | ···················· | 1 |
| B | 1055 | hCoV-19/England/CAMB-7AD28/2020\|EPI_ISL_433868\|202 | ···················· | ···················· | ···················· | ···················· | ···················· | ···················· | |
| B | 1056 | hCoV-19/England/CAMB-7AD46/2020\|EPI_ISL_433870\|202 | ···················· | ···················· | ···················· | ···················· | ···················· | ···················· | |
| B | 1057 | hCoV-19/England/CAMB-7AD55/2020\|EPI_ISL_433871\|202 | ···················· | ···················· | ···················· | ···················· | ···················· | ···················· | |
| B | 1058 | hCoV-19/England/CAMB-7AD64/2020\|EPI_ISL_433872\|202 | ···················· | ···················· | ···················· | ···················· | ···················· | ···················· | |
| B | 1059 | hCoV-19/England/CAMB-7AD82/2020\|EPI_ISL_433873\|202 | ···················· | ···················· | ···················· | ···················· | ···················· | ···················· | |
| B | 1060 | hCoV-19/England/CAMB-7ADA0/2020\|EPI_ISL_433875\|202 | ···················· | ···················· | ···················· | ···················· | ···················· | ···················· | |
| B | 1061 | hCoV-19/England/CAMB-7ADBF/2020\|EPI_ISL_433876\|202 | ···················· | ···················· | ···················· | ···················· | ···················· | ···················· | |
| B | 1062 | hCoV-19/England/CAMB-7ADDD/2020\|EPI_ISL_433877\|202 | ···················· | ···················· | ···················· | ···················· | ···················· | ···················· | |
| B | 1063 | hCoV-19/England/CAMB-7ADEC/2020\|EPI_ISL_433878\|202 | ···················· | ···················· | ···················· | ···················· | ···················· | ···················· | |
| B | 1064 | hCoV-19/England/CAMB-7AE07/2020\|EPI_ISL_433880\|202 | ···················· | ···················· | ···················· | ···················· | ···················· | ···················· | |
| B | 1065 | hCoV-19/England/CAMB-7AE34/2020\|EPI_ISL_433882\|202 | ···················· | ···················· | ···················· | ···················· | ···················· | ···················· | |
| B | 1066 | hCoV-19/England/CAMB-7AE43/2020\|EPI_ISL_433883\|202 | ···················· | ···················· | ···················· | ···················· | ···················· | ···················· | |
| B | 1067 | hCoV-19/England/CAMB-7AE52/2020\|EPI_ISL_433884\|202 | ···················· | ···················· | ···················· | ···················· | ···················· | ···················· | |
| B | 1068 | hCoV-19/England/CAMB-7AE61/2020\|EPI_ISL_433885\|202 | ···················· | ···················· | ···················· | ···················· | ···················· | ···················· | |
| B | 1069 | hCoV-19/USA/TX-HMH0451/2020\|EPI_ISL_435030\|2020-04 | ···················· | ···················· | ···················· | ···················· | ···················· | ···················· | |
| B | 1070 | hCoV-19/USA/TX-HMH0452/2020\|EPI_ISL_435031\|2020-04 | ···················· | ···················· | ···················· | ···················· | ···················· | ···················· | |
| B | 1071 | hCoV-19/DRC/1516/2020\|EPI_ISL_435032\|2020-04-09 | ···················· | ···················· | ···················· | ···················· | ···················· | ···················· | |
| B | 1072 | hCoV-19/DRC/1324/2020\|EPI_ISL_435033\|2020-04-07 | ···················· | ···················· | ···················· | ···················· | ···················· | ···················· | |
| B | 1073 | hCoV-19/USA/LA-EVTL01/2020\|EPI_ISL_435034\|2020-04- | ···················· | ···················· | ···················· | ···················· | ···················· | ···················· | |
| B | 1074 | hCoV-19/USA/LA-EVTL02/2020\|EPI_ISL_435035\|2020-04- | ···················· | ···················· | ···················· | ···················· | ···················· | ···················· | |
| B | 1075 | hCoV-19/USA/LA-EVTL03/2020\|EPI_ISL_435036\|2020-04- | ···················· | ···················· | ···················· | ···················· | ···················· | ···················· | |
| B | 1076 | hCoV-19/USA/LA-EVTL04/2020\|EPI_ISL_435037\|2020-04- | ···················· | ···················· | ···················· | ···················· | ···················· | ···················· | |
| B | 1077 | hCoV-19/USA/LA-EVTL05/2020\|EPI_ISL_435038\|2020-04- | ···················· | ···················· | ···················· | ···················· | ···················· | ···················· | |
| B | 1078 | hCoV-19/USA/LA-EVTL06/2020\|EPI_ISL_435039\|2020-04- | ···················· | ···················· | ···················· | ···················· | ···················· | ···················· | |
| B | 1079 | hCoV-19/USA/LA-EVTL07/2020\|EPI_ISL_435040\|2020-04- | ···················· | ···················· | ···················· | ···················· | ···················· | ···················· | |
| B | 1080 | hCoV-19/USA/LA-EVTL08/2020\|EPI_ISL_435041\|2020-04- | ···················· | ···················· | ···················· | ···················· | ···················· | ···················· | |
| B | 1081 | hCoV-19/USA/LA-EVTL09/2020\|EPI_ISL_435042\|2020-04- | ···················· | ···················· | ···················· | ···················· | ···················· | ···················· | |
| B | 1082 | hCoV-19/USA/LA-EVTL010/2020\|EPI_ISL_435043\|2020-04 | ···················· | ···················· | ···················· | ···················· | ···················· | ···················· | |
| B | 1083 | hCoV-19/USA/LA-EVTL011/2020\|EPI_ISL_435044\|2020-04 | ···················· | ···················· | ···················· | ···················· | ···················· | ···················· | |
| B | 1084 | hCoV-19/Kazakhstan/NCB-1/2020\|EPI_ISL_435045\|2020- | ···················· | ···················· | ···················· | ···················· | ···················· | ···················· | |
| B | 1085 | hCoV-19/Kazakhstan/NCB-2/2020\|EPI_ISL_435046\|2020- | ···················· | ···················· | ···················· | ···················· | ···················· | ···················· | |
| B | 1086 | hCoV-19/Kazakhstan/NCB-3/2020\|EPI_ISL_435047\|2020- | ···················· | ···················· | ···················· | ···················· | ···················· | ···················· | |
| B | 1087 | hCoV-19/Kazakhstan/NCB-5/2020\|EPI_ISL_435048\|2020- | ···················· | ···················· | ···················· | ···················· | ···················· | ···················· | |
| B | 1088 | hCoV-19/India/GJ-GBRC2/2020\|EPI_ISL_435049\|2020-04 | ···················· | ···················· | ···················· | ···················· | ···················· | ···················· | |
| B | 1089 | hCoV-19/Spain/VC-FISABIO-206/2020\|EPI_ISL_436301\|2 | ···················· | ···················· | ···················· | ···················· | ···················· | ···················· | |
| B | 1091 | hCoV-19/Spain/VC-FISABIO-209/2020\|EPI_ISL_436304\|2 | ···················· | ···················· | ···················· | ···················· | ···················· | ···················· | |
| B | 1092 | hCoV-19/Spain/VC-FISABIO-212/2020\|EPI_ISL_436307\|2 | ···················· | ···················· | ···················· | ···················· | ···················· | ···················· | |
| B | 1094 | hCoV-19/Spain/VC-FISABIO-216/2020\|EPI_ISL_436311\|2 | ···················· | ···················· | ···················· | ···················· | ···················· | ···················· | |
| B | 1095 | hCoV-19/Spain/VC-FISABIO-219/2020\|EPI_ISL_436314\|2 | ···················· | ···················· | ···················· | ···················· | ···················· | ···················· | |
| B | 1096 | hCoV-19/Spain/VC-FISABIO-221/2020\|EPI_ISL_436316\|2 | ···················· | ···················· | ···················· | ···················· | ···················· | ···················· | |
| B | 1097 | hCoV-19/Spain/VC-FISABIO-266/2020\|EPI_ISL_436361\|2 | ···················· | ···················· | ···················· | ···················· | ···················· | ···················· | |
| B | 1102 | hCoV-19/Spain/VC-FISABIO-274/2020\|EPI_ISL_436369\|2 | ···················· | ···················· | ···················· | ···················· | ···················· | NNN················· | 1 |
| B | 1103 | hCoV-19/Spain/VC-FISABIO-276/2020\|EPI_ISL_436371\|2 | ···················· | ···················· | ···················· | ···················· | ··N················· | ···················· | |
| B | 1107 | hCoV-19/Spain/VC-FISABIO-282/2020\|EPI_ISL_436377\|2 | ···················· | ···················· | ···················· | ···················· | ···················· | ···················· | |
| B | 1109 | hCoV-19/Spain/VC-FISABIO-284/2020\|EPI_ISL_436379\|2 | ···················· | ···················· | ···················· | ···················· | ···················· | ···················· | |
| B | 1110 | hCoV-19/USA/CA-SR0177/2020\|EPI_ISL_437600\|2020-03- | ···················· | ···················· | ···················· | ···················· | ···················· | ···················· | |
| B | 1111 | hCoV-19/Japan/Donner1/2020\|EPI_ISL_437601\|2020-04- | ···················· | ···················· | ···················· | ···················· | ···················· | ···················· | |
| B | 1112 | hCoV-19/Thailand/SI206917-NT/2020\|EPI_ISL_437602\|2 | ···················· | ···················· | ···················· | ···················· | ···················· | ···················· | |
| B | 1113 | hCoV-19/Thailand/SI206587-NT/2020\|EPI_ISL_437603\|2 | ···················· | ···················· | ···················· | ···················· | ···················· | ···················· | |
| B | 1114 | hCoV-19/Thailand/SI206377-NST/2020\|EPI_ISL_437604\| | ···················· | ···················· | ···················· | ···················· | ···················· | ···················· | |
| B | 1115 | hCoV-19/Thailand/SI205929-NT/2020\|EPI_ISL_437605\|2 | ···················· | ···················· | ···················· | ···················· | ···················· | ···················· | |
| B | 1116 | hCoV-19/Thailand/SI205529-NT/2020\|EPI_ISL_437606\|2 | ···················· | ···················· | ···················· | ···················· | ···················· | ···················· | |
| B | 1117 | hCoV-19/Thailand/SI205457-NST/2020\|EPI_ISL_437607\| | ···················· | ···················· | ···················· | ···················· | ···················· | ···················· | |
| B | 1118 | hCoV-19/Thailand/SI204713-NT/2020\|EPI_ISL_437608\|2 | ···················· | ···················· | ···················· | ···················· | ···················· | ···················· | |
| B | 1119 | hCoV-19/Thailand/SI204514-NT/2020\|EPI_ISL_437609\|2 | ···················· | ···················· | ···················· | ···················· | ···················· | ···················· | |
| B | 1120 | hCoV-19/Thailand/SI204512-NT/2020\|EPI_ISL_437610\|2 | ···················· | ···················· | ···················· | ···················· | ···················· | ···················· | |
| B | 1121 | hCoV-19/Thailand/SI204487-NT/2020\|EPI_ISL_437611\|2 | ···················· | ···················· | ···················· | ···················· | ···················· | ···················· | |
| B | 1122 | hCoV-19/Thailand/SI203285-NT/2020\|EPI_ISL_437612\|2 | ···················· | ···················· | ···················· | ···················· | ···················· | ···················· | |
| B | 1123 | hCoV-19/Thailand/SI202872-NT/2020\|EPI_ISL_437613\|2 | ···················· | ···················· | ···················· | ···················· | ···················· | ···················· | |
| B | 1124 | hCoV-19/Thailand/SI202769-NT/2020\|EPI_ISL_437614\|2 | ···················· | ···················· | ···················· | ···················· | ···················· | ···················· | |
| B | 1125 | hCoV-19/Thailand/SI202620-NT/2020\|EPI_ISL_437615\|2 | ···················· | ···················· | ···················· | ···················· | ···················· | ···················· | |
| B | 1126 | hCoV-19/Thailand/SI202205-NT/2020\|EPI_ISL_437616\|2 | ···················· | ···················· | ···················· | ···················· | ···················· | ···················· | |

| | | | | | | | | | |
|---|---|---|---|---|---|---|---|---|---|
| B | 1127 | hCoV-19/Thailand/SI201712-NT/2020\|EPI_ISL_437617\|2 | .................... | .................... | .................... | .................... | .................... | .................... | |
| B | 1128 | hCoV-19/Thailand/SI200615-NT/2020\|EPI_ISL_437618\|2 | .................... | .................... | .................... | .................... | .................... | .................... | |
| B | 1129 | hCoV-19/Thailand/SI200478-NST/2020\|EPI_ISL_437619\| | .................... | .................... | .................... | .................... | .................... | .................... | |
| B | 1130 | hCoV-19/Thailand/SI200468-NT/2020\|EPI_ISL_437620\|2 | .................... | .................... | .................... | .................... | .................... | .................... | |
| B | 1131 | hCoV-19/Thailand/SI200433-NT/2020\|EPI_ISL_437621\|2 | .................... | .................... | .................... | .................... | .................... | .................... | |
| B | 1132 | hCoV-19/Thailand/SI200383-NT/2020\|EPI_ISL_437622\|2 | .................... | .................... | .................... | .................... | .................... | .................... | |
| B | 1133 | hCoV-19/Thailand/SI200040-NT/2020\|EPI_ISL_437623\|2 | .................... | .................... | .................... | .................... | .................... | .................... | |
| B | 1134 | hCoV-19/Thailand/SI200893-NT/2020\|EPI_ISL_437624\|2 | .................... | .................... | .................... | .................... | .................... | .................... | |
| B | 1135 | hCoV-19/Poland/Wro-02/2020\|EPI_ISL_437625\|2020-03- | .................... | .................... | .................... | .................... | .................... | .................... | |
| B | 1136 | hCoV-19/India/TG-GMC-KP1125/2020\|EPI_ISL_437626\|20 | .................... | .................... | .................... | .................... | .................... | .................... | |
| B | 1137 | hCoV-19/Denmark/ALAB-SSI-1008/2020\|EPI_ISL_437628\| | .................... | .................... | .................... | .................... | .................... | .................... | |
| B | 1138 | hCoV-19/Denmark/ALAB-SSI-1033/2020\|EPI_ISL_437629\| | .................... | .................... | .................... | .................... | .................... | .................... | |
| B | 1139 | hCoV-19/Denmark/ALAB-SSI-1046/2020\|EPI_ISL_437630\| | .................... | .................... | .................... | .................... | .................... | .................... | |
| B | 1140 | hCoV-19/Denmark/ALAB-SSI-1065/2020\|EPI_ISL_437631\| | .................... | .................... | .................... | .................... | .................... | .................... | |
| B | 1141 | hCoV-19/Denmark/ALAB-SSI-1086/2020\|EPI_ISL_437632\| | .................... | .................... | .................... | .................... | .................... | .................... | |
| B | 1142 | hCoV-19/Denmark/ALAB-SSI-1096/2020\|EPI_ISL_437633\| | .................... | .................... | .................... | .................... | .................... | .................... | |
| B | 1143 | hCoV-19/Denmark/ALAB-SSI-1100/2020\|EPI_ISL_437634\| | .................... | .................... | .................... | .................... | .................... | .................... | |
| B | 1144 | hCoV-19/Denmark/ALAB-SSI-1102/2020\|EPI_ISL_437635\| | .................... | .................... | .................... | .................... | .................... | .................... | |
| B | 1145 | hCoV-19/Denmark/ALAB-SSI-1192/2020\|EPI_ISL_437636\| | .................... | .................... | .................... | .................... | .................... | .................... | |
| B | 1146 | hCoV-19/Denmark/ALAB-SSI-1230/2020\|EPI_ISL_437637\| | .................... | .................... | .................... | .................... | .................... | .................... | |
| B | 1147 | hCoV-19/Denmark/ALAB-SSI-1280/2020\|EPI_ISL_437639\| | .................... | .................... | .................... | .................... | .................... | .................... | |
| B | 1148 | hCoV-19/Denmark/ALAB-SSI-1299/2020\|EPI_ISL_437640\| | .................... | .................... | .................... | .................... | .................... | .................... | |
| B | 1149 | hCoV-19/Denmark/ALAB-SSI-133/2020\|EPI_ISL_437642\|2 | .................... | .................... | .................... | .................... | .................... | .................... | |
| B | 1150 | hCoV-19/Denmark/ALAB-SSI-1334/2020\|EPI_ISL_437643\| | .................... | .................... | .................... | .................... | .................... | .................... | |
| B | 1151 | hCoV-19/Denmark/ALAB-SSI-1337/2020\|EPI_ISL_437644\| | .................... | .................... | .................... | .................... | .................... | .................... | |
| B | 1152 | hCoV-19/Denmark/ALAB-SSI-1348/2020\|EPI_ISL_437645\| | .................... | .................... | .................... | .................... | .................... | .................... | |
| B | 1153 | hCoV-19/Denmark/ALAB-SSI-1351/2020\|EPI_ISL_437646\| | .................... | .................... | .................... | .................... | .................... | .................... | |
| B | 1154 | hCoV-19/Denmark/ALAB-SSI-135/2020\|EPI_ISL_437647\|2 | .................... | .................... | .................... | .................... | .................... | .................... | |
| B | 1155 | hCoV-19/France/HDF-CIBU200132/2020\|EPI_ISL_437690\| | .................... | .................... | .................... | .................... | .................... | .................... | |
| B | 1156 | hCoV-19/Saudi Arabia/KAUST-Makkah124/2020\|EPI_ISL_ | .................... | .................... | .................... | .................... | .................... | .................... | |
| B | 1157 | hCoV-19/Saudi Arabia/KAUST-Makkah126/2020\|EPI_ISL_ | .................... | .................... | .................... | .................... | .................... | .................... | |
| B | 1158 | hCoV-19/Saudi Arabia/KAUST-Makkah138/2020\|EPI_ISL_ | .................... | .................... | .................... | .................... | .................... | .................... | |
| B | 1159 | hCoV-19/Saudi Arabia/KAUST-Makkah139/2020\|EPI_ISL_ | .................... | .................... | .................... | .................... | .................... | .................... | |
| B | 1160 | hCoV-19/Saudi Arabia/KAUST-Makkah142/2020\|EPI_ISL_ | .................... | .................... | .................... | .................... | .................... | .................... | |
| B | 1161 | hCoV-19/Saudi Arabia/KAUST-Makkah156/2020\|EPI_ISL_ | .................... | .................... | .................... | .................... | .................... | .................... | |
| B | 1162 | hCoV-19/Saudi Arabia/KAUST-Makkah163/2020\|EPI_ISL_ | .................... | .................... | .................... | .................... | .................... | .................... | |
| B | 1163 | hCoV-19/Saudi Arabia/KAUST-Makkah166/2020\|EPI_ISL_ | .................... | .................... | .................... | .................... | .................... | .................... | |
| B | 1164 | hCoV-19/Saudi Arabia/KAUST-Makkah173/2020\|EPI_ISL_ | .................... | .................... | .................... | .................... | .................... | .................... | |
| B | 1165 | hCoV-19/Scotland/CVR2403/2020\|EPI_ISL_438900\|2020- | .................... | .................... | .................... | .................... | .................... | .................... | |
| B | 1166 | hCoV-19/Scotland/CVR2404/2020\|EPI_ISL_438901\|2020- | .................... | .................... | .................... | .................... | .................... | .................... | |
| B | 1167 | hCoV-19/Scotland/CVR2411/2020\|EPI_ISL_438902\|2020- | .................... | ...................A | .................... | .................... | .................... | .................... | 1 |
| B | 1168 | hCoV-19/Scotland/CVR2413/2020\|EPI_ISL_438903\|2020- | .................... | .................... | .................... | .................... | .................... | .................... | |
| B | 1169 | hCoV-19/Scotland/CVR2437/2020\|EPI_ISL_438905\|2020- | .................... | .................... | .................... | .................... | .................... | .................... | |
| B | 1170 | hCoV-19/Scotland/CVR2446/2020\|EPI_ISL_438907\|2020- | .................... | .................... | .................... | .................... | .................... | .................... | |
| B | 1171 | hCoV-19/Scotland/CVR2447/2020\|EPI_ISL_438908\|2020- | .................... | .................... | .................... | .................... | .................... | .................... | |
| B | 1172 | hCoV-19/Scotland/CVR2450/2020\|EPI_ISL_438910\|2020- | .................... | ...................A | .................... | .................... | .................... | .................... | 1 |
| B | 1173 | hCoV-19/Scotland/CVR2468/2020\|EPI_ISL_438912\|2020- | .................... | .................... | .................... | .................... | .................... | .................... | |
| B | 1174 | hCoV-19/Scotland/CVR247/2020\|EPI_ISL_438913\|2020-0 | .................... | .................... | .................... | .................... | .................... | .................... | |
| B | 1175 | hCoV-19/Scotland/CVR249/2020\|EPI_ISL_438914\|2020-0 | .................... | .................... | .................... | .................... | .................... | .................... | |
| B | 1176 | hCoV-19/Scotland/CVR2492/2020\|EPI_ISL_438915\|2020- | .................... | .................... | .................... | .................... | .................... | .................... | |
| B | 1177 | hCoV-19/Scotland/CVR2496/2020\|EPI_ISL_438916\|2020- | .................... | .................... | .................... | .................... | .................... | .................... | |
| B | 1179 | hCoV-19/Scotland/CVR2512/2020\|EPI_ISL_438920\|2020- | .................... | .................... | .................... | .................... | .................... | .................... | |
| B | 1180 | hCoV-19/Scotland/CVR2515/2020\|EPI_ISL_438921\|2020- | .................... | ...................A | .................... | .................... | .................... | .................... | 1 |
| B | 1181 | hCoV-19/Scotland/CVR2521/2020\|EPI_ISL_438923\|2020- | .................... | .................... | .................... | .................... | .................... | .................... | |
| B | 1182 | hCoV-19/Scotland/CVR2536/2020\|EPI_ISL_438925\|2020- | .................... | .................... | .................... | .................... | .................... | .................... | |
| B | 1183 | hCoV-19/Scotland/CVR255/2020\|EPI_ISL_438926\|2020-0 | .................... | .................... | .................... | .................... | .................... | .................... | |
| B | 1185 | hCoV-19/Scotland/CVR2576/2020\|EPI_ISL_438929\|2020- | .................... | ...................A | .................... | .................... | .................... | .................... | 1 |
| B | 1186 | hCoV-19/Scotland/CVR258/2020\|EPI_ISL_438930\|2020-0 | .................... | .................... | .................... | .................... | .................... | .................... | |
| B | 1187 | hCoV-19/Scotland/CVR2611/2020\|EPI_ISL_438931\|2020- | .................... | .................... | .................... | .................... | .................... | .................... | |
| B | 1188 | hCoV-19/Scotland/CVR263/2020\|EPI_ISL_438933\|2020-0 | .................... | .................... | .................... | .................... | .................... | .................... | |
| B | 1189 | hCoV-19/Scotland/CVR2738/2020\|EPI_ISL_438937\|2020- | .................... | ...................A | .................... | .................... | .................... | .................... | 1 |
| B | 1190 | hCoV-19/Scotland/CVR2743/2020\|EPI_ISL_438941\|2020- | .................... | .................... | .................... | .................... | .................... | .................... | |
| B | 1191 | hCoV-19/Scotland/CVR2746/2020\|EPI_ISL_438944\|2020- | .................... | .................... | .................... | .................... | .................... | .................... | |
| B | 1192 | hCoV-19/Japan/Donner2/2020\|EPI_ISL_438947\|2020-04- | .................... | .................... | .................... | .................... | .................... | .................... | |
| B | 1195 | hCoV-19/Japan/Donner6/2020\|EPI_ISL_438951\|2020-04- | .................... | .................... | .................... | .................... | .................... | .................... | |
| B | 1197 | hCoV-19/Japan/Donner8/2020\|EPI_ISL_438953\|2020-04- | .................... | .................... | .................... | .................... | .................... | .................... | |
| B | 1200 | hCoV-19/Japan/Donner12/2020\|EPI_ISL_438956\|2020-04 | .................... | .................... | .................... | .................... | .................... | .................... | |
| B | 1203 | hCoV-19/Japan/Donner17/2020\|EPI_ISL_438961\|2020-04 | .................... | .................... | .................... | .................... | .................... | .................... | |
| B | 1204 | hCoV-19/Japan/Donner21/2020\|EPI_ISL_438965\|2020-04 | .................... | .................... | .................... | .................... | .................... | .................... | |
| B | 1205 | hCoV-19/Japan/Donner22/2020\|EPI_ISL_438966\|2020-04 | .................... | .................... | .................... | .................... | .................... | .................... | |
| B | 1206 | hCoV-19/Japan/Donner23/2020\|EPI_ISL_438967\|2020-04 | .................... | .................... | .................... | .................... | .................... | .................... | |
| B | 1208 | hCoV-19/Japan/Donner26/2020\|EPI_ISL_438970\|2020-03 | .................... | .................... | .................... | .................... | .................... | .................... | |
| B | 1209 | hCoV-19/Japan/Donner27/2020\|EPI_ISL_438971\|2020-04 | .................... | .................... | .................... | .................... | .................... | .................... | |
| B | 1210 | hCoV-19/Japan/Donner28/2020\|EPI_ISL_438972\|2020-03 | .................... | .................... | .................... | .................... | .................... | .................... | |
| B | 1211 | hCoV-19/Scotland/CVR2749/2020\|EPI_ISL_438973\|2020- | .................... | .................... | .................... | .................... | .................... | .................... | |
| B | 1212 | hCoV-19/Scotland/CVR2750/2020\|EPI_ISL_438974\|2020- | .................... | .................... | .................... | .................... | .................... | .................... | |
| B | 1213 | hCoV-19/Scotland/CVR2751/2020\|EPI_ISL_438975\|2020- | .................... | .................... | .................... | .................... | .................... | .................... | |
| B | 1214 | hCoV-19/Scotland/CVR2757/2020\|EPI_ISL_438976\|2020- | .................... | .................... | .................... | .................... | .................... | .................... | |

| | | | | | | | | |
|---|---|---|---|---|---|---|---|---|
| B | 1215 | hCoV-19/Scotland/CVR2764/2020\|EPI_ISL_438977\|2020- | ···················· | ···················· | ···················· | ···················· | ···················· | ···················· |
| B | 1216 | hCoV-19/Scotland/CVR2770/2020\|EPI_ISL_438978\|2020- | ···················· | ···················· | ···················· | ···················· | ···················· | ···················· |
| B | 1217 | hCoV-19/England/CAMB-7E085/2020\|EPI_ISL_442158\|202 | ···················· | ···················· | ···················· | ···················· | ···················· | ···················· |
| B | 1218 | hCoV-19/England/CAMB-7BB96/2020\|EPI_ISL_442160\|202 | ···················· | ···················· | ···················· | ···················· | ···················· | ···················· |
| B | 1219 | hCoV-19/England/CAMB-7DE22/2020\|EPI_ISL_442161\|202 | ···················· | ·A·················· | ···················· | ···················· | ···················· | ···················· |
| B | 1220 | hCoV-19/England/CAMB-7BDFA/2020\|EPI_ISL_442163\|202 | ···················· | ···················· | ···················· | ···················· | ···················· | ···················· |
| B | 1221 | hCoV-19/England/CAMB-7CE23/2020\|EPI_ISL_442164\|202 | ···················· | ···················· | ···················· | ···················· | ···················· | ···················· |
| B | 1222 | hCoV-19/England/CAMB-7BA5D/2020\|EPI_ISL_442165\|202 | ···················· | ···················· | ···················· | ···················· | ···················· | ···················· |
| B | 1223 | hCoV-19/England/CAMB-7DFD4/2020\|EPI_ISL_442167\|202 | ···················· | ·A·················· | ···················· | ···················· | ···················· | ···················· |
| B | 1224 | hCoV-19/England/CAMB-7CFF3/2020\|EPI_ISL_442168\|202 | ···················· | ···················· | ···················· | ···················· | ···················· | ···················· |
| B | 1225 | hCoV-19/England/CAMB-7E0D0/2020\|EPI_ISL_442169\|202 | ···················· | ···················· | ···················· | ···················· | ···················· | ···················· |
| B | 1226 | hCoV-19/England/CAMB-7B853/2020\|EPI_ISL_442170\|202 | ···················· | ···················· | ···················· | ···················· | ···················· | ···················· |
| B | 1227 | hCoV-19/England/CAMB-7CC38/2020\|EPI_ISL_442171\|202 | ···················· | ···················· | ···················· | ···················· | ···················· | ···················· |
| B | 1228 | hCoV-19/England/CAMB-7DDAD/2020\|EPI_ISL_442172\|202 | ···················· | ···················· | ···················· | ···················· | ···················· | ···················· |
| B | 1229 | hCoV-19/England/CAMB-7B8F9/2020\|EPI_ISL_442174\|202 | ···················· | ···················· | ···················· | ···················· | ···················· | ···················· |
| B | 1230 | hCoV-19/England/CAMB-7DD8F/2020\|EPI_ISL_442175\|202 | ···················· | ···················· | ···················· | ···················· | ···················· | ···················· |
| B | 1231 | hCoV-19/England/CAMB-7BC57/2020\|EPI_ISL_442176\|202 | ···················· | ···················· | ···················· | ···················· | ···················· | ···················· |
| B | 1232 | hCoV-19/England/CAMB-7E0C1/2020\|EPI_ISL_442179\|202 | ···················· | ···················· | ···················· | ···················· | ···················· | ···················· |
| B | 1233 | hCoV-19/England/CAMB-7BBF0/2020\|EPI_ISL_442180\|202 | ···················· | ···················· | ···················· | ···················· | ···················· | ···················· |
| B | 1234 | hCoV-19/England/CAMB-7BC2A/2020\|EPI_ISL_442182\|202 | ···················· | ···················· | ···················· | ···················· | ···················· | ···················· |
| B | 1235 | hCoV-19/England/CAMB-7D00E/2020\|EPI_ISL_442183\|202 | ···················· | ···················· | ···················· | ···················· | ···················· | ···················· |
| B | 1236 | hCoV-19/England/CAMB-7C8F8/2020\|EPI_ISL_442184\|202 | ···················· | ···················· | ···················· | ···················· | ···················· | ···················· |
| B | 1237 | hCoV-19/England/CAMB-7BD36/2020\|EPI_ISL_442186\|202 | ···················· | ···················· | ···················· | ···················· | ···················· | ···················· |
| B | 1238 | hCoV-19/England/CAMB-7CA6B/2020\|EPI_ISL_442187\|202 | ···················· | ···················· | ···················· | ···················· | ···················· | ···················· |
| B | 1239 | hCoV-19/England/CAMB-7CB95/2020\|EPI_ISL_442188\|202 | ···················· | ···················· | ···················· | ···················· | ···················· | ···················· |
| B | 1240 | hCoV-19/England/CAMB-7D02C/2020\|EPI_ISL_442189\|202 | ···················· | ···················· | ···················· | ···················· | ···················· | ···················· |
| B | 1241 | hCoV-19/England/CAMB-7C95F/2020\|EPI_ISL_442190\|202 | ···················· | ···················· | ···················· | ···················· | ···················· | ···················· |
| B | 1242 | hCoV-19/England/CAMB-7C97D/2020\|EPI_ISL_442192\|202 | ···················· | ···················· | ···················· | ···················· | ···················· | ···················· |
| B | 1243 | hCoV-19/England/CAMB-7BCC0/2020\|EPI_ISL_442193\|202 | ···················· | ···················· | ···················· | ···················· | ···················· | ···················· |
| B | 1244 | hCoV-19/England/CAMB-7E058/2020\|EPI_ISL_442194\|202 | ···················· | ···················· | ···················· | ···················· | ···················· | ···················· |
| B | 1245 | hCoV-19/England/CAMB-7E0EF/2020\|EPI_ISL_442196\|202 | ···················· | ···················· | ···················· | ···················· | ···················· | ···················· |
| B | 1246 | hCoV-19/England/CAMB-7CB59/2020\|EPI_ISL_442197\|202 | ···················· | ···················· | ···················· | ···················· | ···················· | ···················· |
| B | 1247 | hCoV-19/England/CAMB-7BCA2/2020\|EPI_ISL_442198\|202 | ···················· | ···················· | ···················· | ···················· | ···················· | ···················· |
| B | 1248 | hCoV-19/England/CAMB-7CA4D/2020\|EPI_ISL_442199\|202 | ···················· | ···················· | ···················· | ···················· | ···················· | ···················· |
| B | 1249 | hCoV-19/Singapore/108/2020\|EPI_ISL_443200\|2020-03- | ···················· | ···················· | ···················· | ···················· | ···················· | ···················· |
| B | 1250 | hCoV-19/Singapore/109/2020\|EPI_ISL_443201\|2020-03- | ···················· | ···················· | ···················· | ···················· | ···················· | ···················· |
| B | 1251 | hCoV-19/Singapore/110/2020\|EPI_ISL_443202\|2020-03- | ···················· | ···················· | ···················· | ···················· | ···················· | ···················· |
| B | 1252 | hCoV-19/England/CAMB-8430B/2020\|EPI_ISL_443489\|202 | ···················· | ···················· | ···················· | ···················· | ···················· | ···················· |
| B | 1253 | hCoV-19/England/CAMB-831B7/2020\|EPI_ISL_443490\|202 | ···················· | ···················· | ···················· | ···················· | ···················· | ···················· |
| B | 1254 | hCoV-19/England/CAMB-8282C/2020\|EPI_ISL_443493\|202 | ···················· | ···················· | ···················· | ···················· | ···················· | ···················· |
| B | 1255 | hCoV-19/England/CAMB-836C7/2020\|EPI_ISL_443494\|202 | ···················· | ···················· | ···················· | ···················· | ···················· | ···················· |
| B | 1256 | hCoV-19/England/CAMB-82895/2020\|EPI_ISL_443495\|202 | ···················· | ···················· | ···················· | ···················· | ···················· | ···················· |
| B | 1257 | hCoV-19/England/CAMB-82789/2020\|EPI_ISL_443496\|202 | ···················· | ···················· | ···················· | ···················· | ···················· | ···················· |
| B | 1258 | hCoV-19/England/CAMB-1AA535/2020\|EPI_ISL_443498\|20 | ···················· | ···················· | ···················· | ···················· | ···················· | ···················· |
| B | 1259 | hCoV-19/USA/WA-QDX-02/2020\|EPI_ISL_444518\|2020-03- | ···················· | ···················· | ···················· | ···················· | ···················· | ···················· |
| B | 1260 | hCoV-19/USA/NY-QDX-01/2020\|EPI_ISL_444519\|2020-03- | ···················· | ···················· | ···················· | ···················· | ···················· | ···················· |
| B | 1265 | hCoV-19/USA/IL-NM014/2020\|EPI_ISL_444525\|2020-03-1 | ···················· | ···················· | ···················· | ···················· | ···················· | ···················· |
| B | 1268 | hCoV-19/USA/IL-NM022/2020\|EPI_ISL_444532\|2020-03-1 | ···················· | ···················· | ···················· | ···················· | ···················· | ···················· |
| B | 1269 | hCoV-19/USA/IL-NM023/2020\|EPI_ISL_444533\|2020-03-1 | ···················· | ···················· | ···················· | ···················· | ···················· | NN·················· |
| B | 1270 | hCoV-19/Wales/PHWC-2BDAE/2020\|EPI_ISL_445846\|2020- | ···················· | ···················· | ···················· | ···················· | ···················· | ···················· |
| B | 1271 | hCoV-19/Wales/PHWC-2BDDB/2020\|EPI_ISL_445849\|2020- | ···················· | ···················· | ···················· | ···················· | ···················· | ···················· |
| B | 1272 | hCoV-19/Wales/PHWC-2BDEA/2020\|EPI_ISL_445850\|2020- | ···················· | ···················· | ···················· | ···················· | ···················· | ···················· |
| B | 1273 | hCoV-19/Wales/PHWC-2BEAB/2020\|EPI_ISL_445862\|2020- | ···················· | ····A··············· | ···················· | ···················· | ···················· | ···················· |
| B | 1274 | hCoV-19/Wales/PHWC-2BEBA/2020\|EPI_ISL_445863\|2020- | ···················· | ···················· | ···················· | ···················· | ···················· | ···················· |
| B | 1275 | hCoV-19/Thailand/Bangkok-0069/2020\|EPI_ISL_447011\| | ···················· | ···················· | ···················· | ···················· | ···················· | C··················· |
| B | 1276 | hCoV-19/Thailand/Bangkok-0074/2020\|EPI_ISL_447015\| | ···················· | ···················· | ···················· | ···················· | ···················· | ···················· |
| B | 1277 | hCoV-19/Thailand/Bangkok-0078/2020\|EPI_ISL_447019\| | ···················· | ···················· | ···················· | ···················· | ···················· | ···················· |
| B | 1278 | hCoV-19/Thailand/Bangkok-0079/2020\|EPI_ISL_447020\| | ···················· | ···················· | ···················· | ···················· | ······K············· | ···················· |
| B | 1279 | hCoV-19/Thailand/Bangkok-0084/2020\|EPI_ISL_447025\| | ···················· | ···················· | ···················· | ···················· | ···················· | ···················· |
| B | 1281 | hCoV-19/Fujian/IM3520003T/2020\|EPI_ISL_431240\|2020 | ···················· | ···················· | ···················· | ···················· | ···················· | ···················· |
| B | 1282 | hCoV-19/England/SHEF-CB0F2/2020\|EPI_ISL_432502\|202 | ···················· | ···················· | ···················· | ···················· | ···················· | ···················· |
| B | 1283 | hCoV-19/England/SHEF-CB317/2020\|EPI_ISL_432505\|202 | ···················· | ···················· | ···················· | ···················· | ···················· | ···················· |
| B | 1284 | hCoV-19/England/SHEF-CB5E4/2020\|EPI_ISL_432510\|202 | ···················· | ···················· | ···················· | ···················· | ···················· | ···················· |
| B | 1285 | hCoV-19/England/SHEF-CBECB/2020\|EPI_ISL_432517\|202 | ···················· | ···················· | ···················· | ···················· | ···················· | ···················· |
| B | 1286 | hCoV-19/England/SHEF-CBF31/2020\|EPI_ISL_432519\|202 | ···················· | ···················· | ···················· | ···················· | ···················· | ···················· |
| B | 1287 | hCoV-19/England/SHEF-CC905/2020\|EPI_ISL_432529\|202 | ···················· | ···················· | ···················· | ···················· | ···················· | ···················· |
| B | 1288 | hCoV-19/England/SHEF-CE89D/2020\|EPI_ISL_432570\|202 | ···················· | ···················· | ···················· | ···················· | ···················· | ···················· |
| B | 1289 | hCoV-19/England/SHEF-CECEC/2020\|EPI_ISL_432578\|202 | ···················· | ···················· | ···················· | ···················· | ···················· | ···················· |
| B | 1290 | hCoV-19/England/SHEF-CED9E/2020\|EPI_ISL_432579\|202 | ···················· | ···················· | ···················· | ···················· | ···················· | ···················· |
| B | 1291 | hCoV-19/England/SHEF-CF340/2020\|EPI_ISL_432588\|202 | ···················· | ···················· | ···················· | ···················· | ···················· | ···················· |
| B | 1292 | hCoV-19/England/CAMB-789AE/2020\|EPI_ISL_433800\|202 | ···················· | ···················· | ···················· | ···················· | ···················· | ···················· |
| B | 1293 | hCoV-19/England/CAMB-789BD/2020\|EPI_ISL_433801\|202 | ···················· | ···················· | ···················· | ···················· | ···················· | ···················· |
| B | 1294 | hCoV-19/England/CAMB-789DB/2020\|EPI_ISL_433803\|202 | ···················· | ···················· | ···················· | ···················· | ···················· | ···················· |
| B | 1295 | hCoV-19/England/CAMB-789F9/2020\|EPI_ISL_433805\|202 | ···················· | ···················· | ···················· | ···················· | ···················· | ···················· |
| B | 1296 | hCoV-19/England/CAMB-78A05/2020\|EPI_ISL_433806\|202 | ···················· | ···G················ | ···················· | ···················· | ···················· | ···················· |
| B | 1297 | hCoV-19/England/CAMB-78A14/2020\|EPI_ISL_433807\|202 | ···················· | ···················· | ···················· | ···················· | ···················· | ···················· |
| B | 1298 | hCoV-19/England/CAMB-78A32/2020\|EPI_ISL_433808\|202 | ···················· | ···················· | ···················· | ···················· | ···················· | ···················· |
| B | 1299 | hCoV-19/England/CAMB-78A50/2020\|EPI_ISL_433809\|202 | ···················· | ···················· | ···················· | ···················· | ···················· | ···················· |

| | | | | | | | |
|---|---|---|---|---|---|---|---|
| B | 1300 hCoV-19/England/CAMB-78A9C/2020\|EPI_ISL_433811\|202 | ···················· | ···················· | ····················· | ···················· | ···················· | ···················· |
| B | 1301 hCoV-19/England/CAMB-78ABA/2020\|EPI_ISL_433812\|202 | ···················· | ···················· | ····················· | ···················· | ···················· | ···················· |
| B | 1302 hCoV-19/England/CAMB-79A04/2020\|EPI_ISL_433813\|202 | ···················· | ···················· | ····················· | ···················· | ···················· | ···················· |
| B | 1303 hCoV-19/England/CAMB-79A22/2020\|EPI_ISL_433814\|202 | ···················· | ···················· | ····················· | ···················· | ···················· | ···················· |
| B | 1304 hCoV-19/England/CAMB-79A40/2020\|EPI_ISL_433815\|202 | ···················· | ···················· | ····················· | ···················· | ···················· | ···················· |
| B | 1305 hCoV-19/England/CAMB-79A5F/2020\|EPI_ISL_433816\|202 | ···················· | ···················· | ····················· | ···················· | ···················· | ···················· |
| B | 1306 hCoV-19/England/CAMB-79A8C/2020\|EPI_ISL_433817\|202 | ···················· | ···················· | ····················· | ···················· | ···················· | ···················· |
| B | 1307 hCoV-19/England/CAMB-79A9B/2020\|EPI_ISL_433818\|202 | ···················· | ···················· | ····················· | ···················· | ···················· | ···················· |
| B | 1308 hCoV-19/England/CAMB-79AB9/2020\|EPI_ISL_433819\|202 | ···················· | ···················· | ····················· | ···················· | ···················· | ···················· |
| B | 1309 hCoV-19/England/CAMB-79AC8/2020\|EPI_ISL_433820\|202 | ···················· | ···················· | ····················· | ···················· | ···················· | ···················· |
| B | 1310 hCoV-19/England/CAMB-79AD7/2020\|EPI_ISL_433821\|202 | ···················· | ···················· | ····················· | ···················· | ···················· | ···················· |
| B | 1311 hCoV-19/England/CAMB-79AF5/2020\|EPI_ISL_433823\|202 | ···················· | ···················· | ····················· | ···················· | ···················· | ···················· |
| B | 1312 hCoV-19/England/CAMB-79B01/2020\|EPI_ISL_433824\|202 | ···················· | ···················· | ····················· | ···················· | ···················· | ···················· |
| B | 1314 hCoV-19/England/CAMB-7A8BE/2020\|EPI_ISL_433826\|202 | ···················· | ···················· | ····················· | ···················· | ···················· | ···················· |
| B | 1315 hCoV-19/England/CAMB-7A8DC/2020\|EPI_ISL_433827\|202 | ···················· | ···················· | ····················· | ···················· | ···················· | ···················· |
| B | 1316 hCoV-19/England/CAMB-7A8FA/2020\|EPI_ISL_433828\|202 | ···················· | ···················· | ····················· | ···················· | ···················· | ···················· |
| B | 1317 hCoV-19/England/CAMB-7AA30/2020\|EPI_ISL_433829\|202 | ···················· | ···················· | ····················· | ···················· | ···················· | ···················· |
| B | 1318 hCoV-19/England/CAMB-7AA8B/2020\|EPI_ISL_433833\|202 | ···················· | ···················· | ····················· | ···················· | ···················· | ···················· |
| B | 1319 hCoV-19/England/CAMB-7AAA9/2020\|EPI_ISL_433834\|202 | ···················· | ···················· | ····················· | ···················· | ···················· | ···················· |
| B | 1320 hCoV-19/England/CAMB-7AAB8/2020\|EPI_ISL_433835\|202 | ···················· | ···················· | ····················· | ···················· | ···················· | ···················· |
| B | 1321 hCoV-19/England/CAMB-7AAD6/2020\|EPI_ISL_433837\|202 | ···················· | ···················· | ····················· | ···················· | ···················· | ···················· |
| B | 1322 hCoV-19/England/CAMB-7AAE5/2020\|EPI_ISL_433838\|202 | ···················· | ···················· | ····················· | ···················· | ···················· | ···················· |
| B | 1323 hCoV-19/England/CAMB-7AAF4/2020\|EPI_ISL_433839\|202 | ···················· | ···················· | ····················· | ···················· | ···················· | ···················· |
| B | 1324 hCoV-19/England/CAMB-7AB00/2020\|EPI_ISL_433840\|202 | ···················· | ···················· | ····················· | ···················· | ···················· | ···················· |
| B | 1325 hCoV-19/England/CAMB-7AB2E/2020\|EPI_ISL_433842\|202 | ···················· | ···················· | ····················· | ···················· | ···················· | ···················· |
| B | 1326 hCoV-19/England/CAMB-7AB3D/2020\|EPI_ISL_433843\|202 | ···················· | ···G················ | ····················· | ···················· | ···················· | ···················· |
| B | 1327 hCoV-19/England/CAMB-7AB79/2020\|EPI_ISL_433844\|202 | ···················· | ···················· | ····················· | ···················· | ···················· | ···················· |
| B | 1328 hCoV-19/England/CAMB-7AB97/2020\|EPI_ISL_433846\|202 | ···················· | ···················· | ····················· | ···················· | ···················· | ···················· |
| B | 1329 hCoV-19/England/CAMB-7ABA6/2020\|EPI_ISL_433847\|202 | ···················· | ···················· | ····················· | ···················· | ···················· | ···················· |
| B | 1330 hCoV-19/England/CAMB-7ABB5/2020\|EPI_ISL_433848\|202 | ···················· | ···················· | ····················· | ···················· | ···················· | ···················· |
| B | 1331 hCoV-19/England/CAMB-7ABC4/2020\|EPI_ISL_433849\|202 | ···················· | ···················· | ····················· | ···················· | ···················· | ···················· |
| B | 1332 hCoV-19/England/CAMB-7ABF1/2020\|EPI_ISL_433851\|202 | ···················· | ···················· | ····················· | ···················· | ···················· | ···················· |
| B | 1333 hCoV-19/England/CAMB-7AC0D/2020\|EPI_ISL_433852\|202 | ···················· | ···················· | ····················· | ···················· | ···················· | ···················· |
| B | 1334 hCoV-19/England/CAMB-7AC2B/2020\|EPI_ISL_433854\|202 | ···················· | ···················· | ····················· | ···················· | ···················· | ···················· |
| B | 1335 hCoV-19/England/CAMB-7AC3A/2020\|EPI_ISL_433855\|202 | ···················· | ···················· | ····················· | ···················· | ···················· | ···················· |
| B | 1336 hCoV-19/England/CAMB-7AC49/2020\|EPI_ISL_433856\|202 | ···················· | ···················· | ····················· | ···················· | ···················· | ···················· |
| B | 1337 hCoV-19/England/CAMB-7AC58/2020\|EPI_ISL_433857\|202 | ···················· | ···················· | ····················· | ···················· | ···················· | ···················· |
| B | 1338 hCoV-19/England/CAMB-7AC67/2020\|EPI_ISL_433858\|202 | ···················· | ···················· | ····················· | ···················· | ···················· | ···················· |
| B | 1339 hCoV-19/England/CAMB-7AC76/2020\|EPI_ISL_433859\|202 | ···················· | ···················· | ····················· | ···················· | ···················· | ···················· |
| B | 1340 hCoV-19/England/CAMB-7AC85/2020\|EPI_ISL_433860\|202 | ···················· | ···················· | ····················· | ···················· | ···················· | ···················· |
| B | 1341 hCoV-19/England/CAMB-7AC94/2020\|EPI_ISL_433861\|202 | ···················· | ···················· | ····················· | ···················· | ···················· | ···················· |
| B | 1342 hCoV-19/England/CAMB-7ACA3/2020\|EPI_ISL_433862\|202 | ···················· | ···················· | ····················· | ···················· | ···················· | ···················· |
| B | 1343 hCoV-19/England/CAMB-7ACB2/2020\|EPI_ISL_433863\|202 | ···················· | ···················· | ····················· | ···················· | ···················· | ···················· |
| B | 1344 hCoV-19/England/CAMB-7AE70/2020\|EPI_ISL_433886\|202 | ···················· | ···················· | ····················· | ···················· | ···················· | ···················· |
| B | 1345 hCoV-19/England/CAMB-7AEAD/2020\|EPI_ISL_433888\|202 | ···················· | ···················· | ····················· | ···················· | ···················· | ···················· |
| B | 1346 hCoV-19/England/CAMB-7AEDA/2020\|EPI_ISL_433890\|202 | ···················· | ···················· | ····················· | ···················· | ···················· | ···················· |
| B | 1347 hCoV-19/England/CAMB-7AEE9/2020\|EPI_ISL_433891\|202 | ···················· | ···················· | ····················· | ···················· | ···················· | ···················· |
| B | 1348 hCoV-19/England/CAMB-7AEF8/2020\|EPI_ISL_433892\|202 | ···················· | ···················· | ····················· | ···················· | ···················· | ···················· |
| B | 1349 hCoV-19/England/CAMB-7AF04/2020\|EPI_ISL_433893\|202 | ···················· | ···················· | ····················· | ···················· | ···················· | ···················· |
| B | 1350 hCoV-19/England/CAMB-7AF13/2020\|EPI_ISL_433894\|202 | ···················· | ···················· | ····················· | ···················· | ···················· | ···················· |
| B | 1351 hCoV-19/England/CAMB-7AF22/2020\|EPI_ISL_433895\|202 | ···················· | ···················· | ····················· | ···················· | ···················· | ···················· |
| B | 1352 hCoV-19/England/CAMB-7AF31/2020\|EPI_ISL_433896\|202 | ···················· | ···G················ | ····················· | ···················· | ···················· | ···················· |
| B | 1353 hCoV-19/England/CAMB-7AF40/2020\|EPI_ISL_433897\|202 | ···················· | ···················· | ····················· | ···················· | ···················· | ···················· |
| B | 1354 hCoV-19/England/CAMB-7AF5F/2020\|EPI_ISL_433898\|202 | ···················· | ···················· | ····················· | ···················· | ···················· | ···················· |
| B | 1355 hCoV-19/England/CAMB-7AF6E/2020\|EPI_ISL_433899\|202 | ···················· | ···················· | ····················· | ···················· | ···················· | ···················· |
| B | 1356 hCoV-19/USA/DC-CDC-9096/2020\|EPI_ISL_436040\|2020-0 | ···················· | ···················· | ····················· | ···················· | ···················· | ···················· |
| B | 1357 hCoV-19/USA/DC-CDC-9092/2020\|EPI_ISL_436041\|2020-0 | ···················· | ···················· | ····················· | ···················· | ···················· | ···················· |
| B | 1358 hCoV-19/USA/DC-CDC-1734/2020\|EPI_ISL_436042\|2020-0 | ···················· | ···················· | ····················· | ···················· | ···················· | ···················· |
| B | 1359 hCoV-19/USA/DC-CDC-1738/2020\|EPI_ISL_436043\|2020-0 | ···················· | ···················· | ····················· | ···················· | ···················· | ···················· |
| B | 1360 hCoV-19/USA/LA-CDC-1894/2020\|EPI_ISL_436044\|2020-0 | ···················· | ···················· | ····················· | ···················· | ···················· | ···················· |
| B | 1361 hCoV-19/USA/VI-CDC-1999/2020\|EPI_ISL_436045\|2020-0 | ···················· | ···················· | ····················· | ···················· | ···················· | ···················· |
| B | 1362 hCoV-19/USA/VI-CDC-1993/2020\|EPI_ISL_436046\|2020-0 | ···················· | ···················· | ····················· | ···················· | ···················· | ···················· |
| B | 1363 hCoV-19/USA/NY-SURV052/2020\|EPI_ISL_436047\|2020-03 | ···················· | ···················· | ····················· | ···················· | ···················· | ···················· |
| B | 1364 hCoV-19/USA/NY-SURV076/2020\|EPI_ISL_436048\|2020-03 | ···················· | ···················· | ····················· | ···················· | ···················· | ···················· |
| B | 1365 hCoV-19/USA/NY-SURV097/2020\|EPI_ISL_436049\|2020-03 | ···················· | ···················· | ····················· | ···················· | ···················· | ···················· |
| B | 1366 hCoV-19/USA/NY-SURV0144/2020\|EPI_ISL_436050\|2020-0 | ···················· | ···················· | ····················· | ···················· | ···················· | ···················· |
| B | 1367 hCoV-19/USA/NY-SURV0162/2020\|EPI_ISL_436051\|2020-0 | ···················· | ···················· | ····················· | ···················· | ···················· | ···················· |
| B | 1368 hCoV-19/USA/NY-SURV0165/2020\|EPI_ISL_436052\|2020-0 | ···················· | ···················· | ····················· | ···················· | ···················· | ···················· |
| B | 1369 hCoV-19/USA/NY-SURV0166/2020\|EPI_ISL_436053\|2020-0 | ···················· | ···················· | ····················· | ···················· | ···················· | ···················· |
| B | 1370 hCoV-19/USA/NY-SURV0168/2020\|EPI_ISL_436054\|2020-0 | ···················· | ···················· | ····················· | ···················· | ···················· | ···················· |
| B | 1371 hCoV-19/USA/NY-SURV0171/2020\|EPI_ISL_436055\|2020-0 | ···················· | ···················· | ····················· | ···················· | ···················· | ···················· |
| B | 1372 hCoV-19/USA/NY-SURV0175/2020\|EPI_ISL_436056\|2020-0 | ···················· | ···················· | ····················· | ···················· | ···················· | ···················· |
| B | 1373 hCoV-19/USA/NY-SURV0183/2020\|EPI_ISL_436057\|2020-0 | ···················· | ···················· | ····················· | ···················· | ···················· | ···················· |
| B | 1374 hCoV-19/USA/NY-SURV0186/2020\|EPI_ISL_436058\|2020-0 | ···················· | ···················· | ····················· | ···················· | ···················· | ···················· |
| B | 1375 hCoV-19/USA/NY-SURV0240/2020\|EPI_ISL_436059\|2020-0 | ···················· | ···················· | ····················· | ···················· | ···················· | ···················· |
| B | 1376 hCoV-19/USA/NY-SURV0302/2020\|EPI_ISL_436060\|2020-0 | ···················· | ···················· | ····················· | ···················· | ···················· | ···················· |
| B | 1377 hCoV-19/USA/NY-SURV0319/2020\|EPI_ISL_436061\|2020-0 | ···················· | ···················· | ····················· | ···················· | ···················· | ···················· |
| B | 1378 hCoV-19/USA/NY-SURV0331/2020\|EPI_ISL_436062\|2020-0 | ···················· | ···················· | ····················· | ···················· | ···················· | ···················· |

| | | | | | | | | | |
|---|---|---|---|---|---|---|---|---|---|
| B | 1379 | hCoV-19/USA/NY-SURV0334/2020\|EPI_ISL_436063\|2020-0 | .................... | .................... | .................... | .................... | .................... | .................... | |
| B | 1380 | hCoV-19/USA/NY-SURV0397/2020\|EPI_ISL_436064\|2020-0 | .................... | .................... | .................... | .................... | .................... | .................... | |
| B | 1381 | hCoV-19/USA/NY-SURV0407/2020\|EPI_ISL_436065\|2020-0 | .................... | .................... | .................... | .................... | .................... | .................... | |
| B | 1382 | hCoV-19/USA/NY-SURV0440/2020\|EPI_ISL_436066\|2020-0 | .................... | .................... | .................... | .................... | .................... | .................... | |
| B | 1383 | hCoV-19/USA/NY-SURV0444/2020\|EPI_ISL_436067\|2020-0 | .................... | .................... | .................... | .................... | .................... | .................... | |
| B | 1384 | hCoV-19/USA/NY-SURV0472/2020\|EPI_ISL_436068\|2020-0 | .................... | .................... | .................... | .................... | .................... | .................... | |
| B | 1385 | hCoV-19/USA/NY-SURV0475/2020\|EPI_ISL_436069\|2020-0 | .................... | .................... | .................... | .................... | .................... | .................... | |
| B | 1386 | hCoV-19/USA/NY-SURV0495/2020\|EPI_ISL_436070\|2020-0 | .................... | .................... | .................... | .................... | .................... | .................... | |
| B | 1387 | hCoV-19/USA/NY-SURV0513/2020\|EPI_ISL_436071\|2020-0 | .................... | .................... | .................... | .................... | .................... | .................... | |
| B | 1388 | hCoV-19/USA/NY-SURV0592/2020\|EPI_ISL_436072\|2020-0 | .................... | .................... | .................... | .................... | .................... | .................... | |
| B | 1389 | hCoV-19/USA/NY-SURV0710/2020\|EPI_ISL_436073\|2020-0 | .................... | .................... | .................... | .................... | .................... | .................... | |
| B | 1390 | hCoV-19/USA/NY-SURV0862/2020\|EPI_ISL_436074\|2020-0 | .................... | .................... | .................... | .................... | .................... | .................... | |
| B | 1391 | hCoV-19/USA/NY-SURV0863/2020\|EPI_ISL_436075\|2020-0 | .................... | .................... | .................... | .................... | .................... | .................... | |
| B | 1392 | hCoV-19/USA/NY-SURV0874/2020\|EPI_ISL_436076\|2020-0 | .................... | .................... | .................... | .................... | .................... | .................... | |
| B | 1393 | hCoV-19/USA/NY-SURV0982/2020\|EPI_ISL_436077\|2020-0 | .................... | .................... | .................... | .................... | .................... | .................... | |
| B | 1394 | hCoV-19/USA/NY-SURV0983/2020\|EPI_ISL_436078\|2020-0 | .................... | .................... | .................... | .................... | .................... | .................... | |
| B | 1395 | hCoV-19/USA/NY-SURV0985/2020\|EPI_ISL_436079\|2020-0 | .................... | .................... | .................... | .................... | .................... | .................... | |
| B | 1396 | hCoV-19/USA/NY-SURV0988/2020\|EPI_ISL_436080\|2020-0 | .................... | .................... | .................... | .................... | .................... | .................... | |
| B | 1397 | hCoV-19/USA/NY-SURV0996/2020\|EPI_ISL_436081\|2020-0 | .................... | .................... | .................... | .................... | .................... | .................... | |
| B | 1398 | hCoV-19/USA/NY-SURV1000/2020\|EPI_ISL_436082\|2020-0 | .................... | .................... | .................... | .................... | .................... | .................... | |
| B | 1399 | hCoV-19/Australia/QLDID935/2020\|EPI_ISL_436097\|202 | .................... | .................... | .................... | .................... | .................... | .................... | |
| B | 1400 | hCoV-19/Australia/QLDID936/2020\|EPI_ISL_436098\|202 | .................... | .................... | .................... | .................... | .................... | .................... | |
| B | 1401 | hCoV-19/Taiwan/TSGH-24/2020\|EPI_ISL_436099\|2020-04 | .................... | .................... | .................... | .................... | .................... | .................... | |
| B | 1402 | hCoV-19/Turkey/HSGM-439/2020\|EPI_ISL_437307\|2020-0 | .................... | .................... | .................... | .................... | .................... | .................... | |
| B | 1403 | hCoV-19/Turkey/HSGM-515/2020\|EPI_ISL_437308\|2020-0 | .................... | .................... | .................... | .................... | .................... | .................... | |
| B | 1404 | hCoV-19/Turkey/HSGM-1049/2020\|EPI_ISL_437309\|2020- | .................... | .................... | .................... | .................... | .................... | .................... | |
| B | 1405 | hCoV-19/Turkey/HSGM-510/2020\|EPI_ISL_437314\|2020-0 | .................... | .................... | .................... | .................... | .................... | .................... | |
| B | 1406 | hCoV-19/Turkey/HSGM-1004/2020\|EPI_ISL_437316\|2020- | .................... | .................... | .................... | .................... | .................... | .................... | |
| B | 1407 | hCoV-19/Turkey/HSGM-1027/2020\|EPI_ISL_437317\|2020- | .................... | .................... | .................... | .................... | .................... | .................... | |
| B | 1408 | hCoV-19/USA/MN-MDH-400/2020\|EPI_ISL_437370\|2020-04 | .................... | .................... | .................... | .................... | .................... | .................... | |
| B | 1409 | hCoV-19/USA/MN-MDH-418/2020\|EPI_ISL_437371\|2020-04 | .................... | .................... | .................... | .................... | .................... | .................... | |
| B | 1410 | hCoV-19/USA/MN-MDH-435/2020\|EPI_ISL_437372\|2020-04 | .................... | .................... | .................... | .................... | .................... | .................... | |
| B | 1411 | hCoV-19/USA/MN-MDH-436/2020\|EPI_ISL_437373\|2020-04 | .................... | .................... | .................... | .................... | .................... | .................... | |
| B | 1412 | hCoV-19/USA/MN-MDH-470/2020\|EPI_ISL_437374\|2020-04 | .................... | .................... | .................... | .................... | .................... | .................... | |
| B | 1413 | hCoV-19/USA/MN-MDH-472/2020\|EPI_ISL_437375\|2020-04 | .................... | ....G............... | .................... | .................... | .................... | .................... | |
| B | 1414 | hCoV-19/USA/MN-MDH-480/2020\|EPI_ISL_437376\|2020-04 | .................... | .................... | .................... | .................... | .................... | .................... | |
| B | 1415 | hCoV-19/USA/MN-MDH-506/2020\|EPI_ISL_437377\|2020-04 | .................... | .................... | .................... | .................... | .................... | .................... | |
| B | 1416 | hCoV-19/USA/MN-MDH-513/2020\|EPI_ISL_437378\|2020-04 | .................... | .................... | .................... | .................... | .................... | .................... | |
| B | 1417 | hCoV-19/USA/MN-MDH-517/2020\|EPI_ISL_437379\|2020-04 | .................... | .................... | .................... | .................... | .................... | .................... | |
| B | 1418 | hCoV-19/USA/MN-MDH-564/2020\|EPI_ISL_437380\|2020-04 | .................... | .................... | .................... | .................... | .................... | .................... | |
| B | 1419 | hCoV-19/USA/MN-MDH-578/2020\|EPI_ISL_437381\|2020-04 | .................... | .................... | .................... | .................... | .................... | .................... | |
| B | 1420 | hCoV-19/USA/MN-MDH-585/2020\|EPI_ISL_437382\|2020-04 | .................... | .................... | ...A................ | .................... | .................... | .................... | |
| B | 1421 | hCoV-19/USA/MN-MDH-621/2020\|EPI_ISL_437383\|2020-04 | .................... | .................... | ...A................ | .................... | .................... | .................... | |
| B | 1422 | hCoV-19/USA/MN-MDH-637/2020\|EPI_ISL_437384\|2020-04 | .................... | ....A............... | .................... | .................... | .................... | .................... | |
| B | 1423 | hCoV-19/USA/MN-MDH-640/2020\|EPI_ISL_437385\|2020-04 | .................... | .................... | .................... | .................... | .................... | .................... | |
| B | 1424 | hCoV-19/USA/MN-MDH-666/2020\|EPI_ISL_437386\|2020-04 | .................... | .................... | .................... | .................... | .................... | .................... | |
| B | 1425 | hCoV-19/USA/VA-DCLS-0171/2020\|EPI_ISL_437387\|2020- | .................... | .................... | .................... | .................... | .................... | .................... | |
| B | 1426 | hCoV-19/USA/VA-DCLS-0172/2020\|EPI_ISL_437388\|2020- | .................... | .................... | .................... | .................... | .................... | .................... | |
| B | 1427 | hCoV-19/USA/VA-DCLS-0173/2020\|EPI_ISL_437389\|2020- | .................... | .................... | .................... | .................... | .................... | .................... | |
| B | 1428 | hCoV-19/USA/VA-DCLS-0174/2020\|EPI_ISL_437390\|2020- | .................... | .................... | .................... | .................... | .................... | .................... | |
| B | 1429 | hCoV-19/USA/VA-DCLS-0175/2020\|EPI_ISL_437391\|2020- | .................... | .................... | .................... | .................... | .................... | .................... | |
| B | 1430 | hCoV-19/USA/VA-DCLS-0176/2020\|EPI_ISL_437392\|2020- | .................... | .................... | .................... | .................... | .................... | .................... | |
| B | 1431 | hCoV-19/USA/VA-DCLS-0177/2020\|EPI_ISL_437393\|2020- | .................... | .................... | .................... | .................... | .................... | .................... | |
| B | 1432 | hCoV-19/USA/VA-DCLS-0178/2020\|EPI_ISL_437394\|2020- | .................... | .................... | .................... | .................... | .................... | .................... | |
| B | 1433 | hCoV-19/USA/VA-DCLS-0179/2020\|EPI_ISL_437395\|2020- | .................... | .................... | .................... | .................... | .................... | .................... | |
| B | 1434 | hCoV-19/USA/VA-DCLS-0180/2020\|EPI_ISL_437396\|2020- | .................... | .................... | .................... | .................... | .................... | .................... | |
| B | 1435 | hCoV-19/USA/VA-DCLS-0181/2020\|EPI_ISL_437397\|2020- | .................... | .................... | .................... | .................... | .................... | .................... | |
| B | 1436 | hCoV-19/USA/VA-DCLS-0182/2020\|EPI_ISL_437398\|2020- | .................... | .................... | .................... | .................... | .................... | .................... | |
| B | 1437 | hCoV-19/USA/VA-DCLS-0183/2020\|EPI_ISL_437399\|2020- | .................... | .................... | .................... | .................... | .................... | .................... | |
| B | 1438 | hCoV-19/England/CAMB-829FC/2020\|EPI_ISL_438645\|202 | .................... | .................... | .................... | .................... | .................... | .................... | |
| B | 1439 | hCoV-19/England/CAMB-82A17/2020\|EPI_ISL_438647\|202 | .................... | .................... | .................... | .................... | .................... | .................... | |
| B | 1440 | hCoV-19/England/BRIS-122210/2020\|EPI_ISL_439969\|20 | .................... | .................... | .................... | .................... | .................... | .................... | |
| B | 1441 | hCoV-19/England/BRIS-12218C/2020\|EPI_ISL_439978\|20 | .................... | .................... | .................... | .................... | .................... | .................... | |
| B | 1442 | hCoV-19/England/BRIS-121F47/2020\|EPI_ISL_439979\|20 | .................... | .................... | .................... | .................... | .................... | .................... | |
| B | 1443 | hCoV-19/England/LOND-D3561/2020\|EPI_ISL_440956\|202 | .................... | .................... | .................... | .................... | .................... | .................... | |
| B | 1444 | hCoV-19/England/LOND-D358F/2020\|EPI_ISL_440957\|202 | .................... | .................... | .................... | .................... | .................... | .................... | |
| B | 1445 | hCoV-19/England/LOND-D359E/2020\|EPI_ISL_440958\|202 | .................... | .................... | .................... | .................... | .................... | .................... | |
| B | 1446 | hCoV-19/Singapore/111/2020\|EPI_ISL_443203\|2020-03- | .................... | .................... | .................... | .................... | .................... | .................... | |
| B | 1447 | hCoV-19/Singapore/112/2020\|EPI_ISL_443204\|2020-03- | .................... | .................... | .................... | .................... | .................... | .................... | |
| B | 1448 | hCoV-19/Singapore/113/2020\|EPI_ISL_443205\|2020-03- | .................... | .................... | .................... | .................... | .................... | .................... | |
| B | 1449 | hCoV-19/Singapore/114/2020\|EPI_ISL_443206\|2020-03- | .................... | .................... | .................... | .................... | .................... | .................... | |
| B | 1450 | hCoV-19/Singapore/115/2020\|EPI_ISL_443207\|2020-03- | .................... | .................... | .................... | .................... | .................... | .................... | |
| B | 1451 | hCoV-19/Singapore/116/2020\|EPI_ISL_443208\|2020-03- | .................... | ...................A | .................... | .................... | .................... | .................... | 1 |
| B | 1452 | hCoV-19/Singapore/117/2020\|EPI_ISL_443209\|2020-03- | .................... | .................... | .................... | .................... | .................... | .................... | |
| B | 1453 | hCoV-19/Singapore/118/2020\|EPI_ISL_443210\|2020-03- | .................... | .................... | .................... | .................... | .................... | .................... | |
| B | 1454 | hCoV-19/Singapore/119/2020\|EPI_ISL_443211\|2020-03- | .................... | .................... | .................... | .................... | .................... | .................... | |
| B | 1455 | hCoV-19/Singapore/120/2020\|EPI_ISL_443212\|2020-04- | .................... | .................... | .................... | .................... | .................... | .................... | |
| B | 1456 | hCoV-19/Singapore/121/2020\|EPI_ISL_443213\|2020-04- | .................... | .................... | .................... | .................... | .................... | .................... | |

| | | | | | | | |
|---|---|---|---|---|---|---|---|
| B | 1457 hCoV-19/Singapore/122/2020\|EPI_ISL_443214\|2020-03- | ···················· | ···················· | ···················· | ···················· | ···················· | ···················· |
| B | 1458 hCoV-19/Singapore/123/2020\|EPI_ISL_443215\|2020-03- | ···················· | ···················· | ···················· | ···················· | ···················· | ···················· |
| B | 1459 hCoV-19/Singapore/124/2020\|EPI_ISL_443216\|2020-03- | ···················· | ···················· | ···················· | ···················· | ···················· | ···················· |
| B | 1460 hCoV-19/Singapore/125/2020\|EPI_ISL_443217\|2020-03- | ···················· | ···················· | ···················· | ···················· | ···················· | ···················· |
| B | 1461 hCoV-19/Singapore/126/2020\|EPI_ISL_443218\|2020-03- | ···················· | ···················· | ···················· | ···················· | ···················· | ···················· |
| B | 1462 hCoV-19/Singapore/127/2020\|EPI_ISL_443219\|2020-03- | ···················· | ···················· | ···················· | ···················· | ···················· | ···················· |
| B | 1463 hCoV-19/Singapore/128/2020\|EPI_ISL_443220\|2020-03- | ···················· | ···················· | ···················· | ···················· | ···················· | ···················· |
| B | 1464 hCoV-19/Singapore/129/2020\|EPI_ISL_443221\|2020-03- | ···················· | ···················· | ···················· | ···················· | ···················· | ···················· |
| B | 1465 hCoV-19/Singapore/130/2020\|EPI_ISL_443222\|2020-03- | ···················· | ···················· | ···················· | ···················· | ···················· | ···················· |
| B | 1466 hCoV-19/Singapore/131/2020\|EPI_ISL_443223\|2020-03- | ···················· | ···················· | ···················· | ···················· | ···················· | ···················· |
| B | 1467 hCoV-19/Singapore/132/2020\|EPI_ISL_443224\|2020-03- | ···················· | ···················· | ···················· | ···················· | ···················· | ···················· |
| B | 1468 hCoV-19/Singapore/133/2020\|EPI_ISL_443225\|2020-03- | ···················· | ···················· | ···················· | ···················· | ···················· | ···················· |
| B | 1469 hCoV-19/Singapore/134/2020\|EPI_ISL_443226\|2020-03- | ···················· | ···················· | ···················· | ···················· | ···················· | ···················· |
| B | 1470 hCoV-19/Singapore/135/2020\|EPI_ISL_443227\|2020-03- | ···················· | ···················· | ···················· | ···················· | ···················· | ···················· |
| B | 1471 hCoV-19/Singapore/136/2020\|EPI_ISL_443228\|2020-03- | ···················· | ···················· | ···················· | ···················· | ···················· | ···················· |
| B | 1472 hCoV-19/Singapore/137/2020\|EPI_ISL_443229\|2020-03- | ···················· | ···················· | ···················· | ···················· | ···················· | ···················· |
| B | 1473 hCoV-19/Singapore/138/2020\|EPI_ISL_443230\|2020-03- | ···················· | ···················· | ···················· | ···················· | ···················· | ···················· |
| B | 1474 hCoV-19/Singapore/139/2020\|EPI_ISL_443231\|2020-03- | ···················· | ···················· | ···················· | ···················· | ···················· | ···················· |
| B | 1475 hCoV-19/Singapore/140/2020\|EPI_ISL_443232\|2020-03- | ···················· | ···················· | ···················· | ···················· | ···················· | ···················· |
| B | 1476 hCoV-19/Singapore/141/2020\|EPI_ISL_443233\|2020-03- | ···················· | ···················· | ···················· | ···················· | ···················· | ···················· |
| B | 1477 hCoV-19/Singapore/142/2020\|EPI_ISL_443234\|2020-03- | ···················· | ···················· | ···················· | ···················· | ···················· | ···················· |
| B | 1478 hCoV-19/Singapore/143/2020\|EPI_ISL_443235\|2020-03- | ···················· | ···················· | ···················· | ···················· | ···················· | ···················· |
| B | 1479 hCoV-19/Singapore/144/2020\|EPI_ISL_443236\|2020-03- | ···················· | ···················· | ···················· | ···················· | ···················· | ···················· |
| B | 1480 hCoV-19/Singapore/145/2020\|EPI_ISL_443237\|2020-03- | ···················· | ···················· | ···················· | ···················· | ···················· | ···················· |
| B | 1481 hCoV-19/Singapore/146/2020\|EPI_ISL_443238\|2020-03- | ···················· | ···················· | ···················· | ···················· | ···················· | ···················· |
| B | 1482 hCoV-19/Singapore/147/2020\|EPI_ISL_443239\|2020-03- | ···················· | ···················· | ···················· | ···················· | ···················· | ···················· |
| B | 1483 hCoV-19/Singapore/148/2020\|EPI_ISL_443240\|2020-03- | ···················· | ···················· | ···················· | ···················· | ···················· | ···················· |
| B | 1484 hCoV-19/Singapore/149/2020\|EPI_ISL_443241\|2020-03- | ···················· | ···················· | ···················· | ···················· | ···················· | ···················· |
| B | 1485 hCoV-19/Singapore/150/2020\|EPI_ISL_443242\|2020-03- | ···················· | ···················· | ···················· | ···················· | ···················· | ···················· |
| B | 1486 hCoV-19/Singapore/151/2020\|EPI_ISL_443243\|2020-03- | ···················· | ···················· | ···················· | ···················· | ···················· | ···················· |
| B | 1487 hCoV-19/Singapore/152/2020\|EPI_ISL_443244\|2020-04- | ···················· | ···················· | ···················· | ···················· | ···················· | ···················· |
| B | 1488 hCoV-19/Singapore/153/2020\|EPI_ISL_443245\|2020-04- | ···················· | ···················· | ···················· | ···················· | ···················· | ···················· |
| B | 1489 hCoV-19/Singapore/154/2020\|EPI_ISL_443246\|2020-04- | ···················· | ···················· | ···················· | ···················· | ···················· | ···················· |
| B | 1490 hCoV-19/Canada/QC_AB2/2020\|EPI_ISL_444500\|2020-04- | ···················· | ···················· | ···················· | ···················· | ···················· | ···················· |
| B | 1503 hCoV-19/USA/IL-NM054/2020\|EPI_ISL_444550\|2020-03-1 | ···················· | ···················· | ···················· | ···················· | ···················· | ···················· |
| B | 1504 hCoV-19/USA/IL-NM056/2020\|EPI_ISL_444552\|2020-03-1 | ···················· | ···················· | ···················· | ···················· | ···················· | ···················· |
| B | 1506 hCoV-19/USA/IL-NM059/2020\|EPI_ISL_444555\|2020-03-1 | ···················· | ···················· | ···················· | ···················· | ···················· | ···················· |
| B | 1507 hCoV-19/USA/IL-NM060/2020\|EPI_ISL_444556\|2020-03-1 | ···················· | ···················· | ···················· | ···················· | ···················· | ···················· |
| B | 1509 hCoV-19/USA/IL-NM065/2020\|EPI_ISL_444560\|2020-03-1 | ···················· | ···················· | ···················· | ···················· | ···················· | ···················· |
| B | 1513 hCoV-19/USA/IL-NM071/2020\|EPI_ISL_444566\|2020-03-1 | ···················· | ···················· | ···················· | ···················· | ···················· | ···················· |
| B | 1514 hCoV-19/USA/IL-NM072/2020\|EPI_ISL_444567\|2020-03-1 | ···················· | ···················· | ···················· | ···················· | ···················· | ···················· |
| B | 1515 hCoV-19/USA/IL-NM073/2020\|EPI_ISL_444568\|2020-03-1 | ···················· | ···················· | ···················· | ···················· | ···················· | ···················· |
| B | 1516 hCoV-19/USA/IL-NM074/2020\|EPI_ISL_444569\|2020-03-1 | ···················· | ···················· | ···················· | ···················· | ···················· | ···················· |
| B | 1517 hCoV-19/USA/IL-NM075/2020\|EPI_ISL_444570\|2020-03-1 | ···················· | ···················· | ···················· | ···················· | ···················· | ···················· |
| B | 1518 hCoV-19/USA/IL-NM079/2020\|EPI_ISL_444571\|2020-03-1 | ···················· | ···················· | ···················· | ···················· | ···················· | ···················· |
| B | 1520 hCoV-19/USA/IL-NM082/2020\|EPI_ISL_444573\|2020-03-1 | ···················· | ···················· | ···················· | ···················· | ···················· | ···················· |
| B | 1521 hCoV-19/USA/IL-NM083/2020\|EPI_ISL_444574\|2020-03-1 | ···················· | ···················· | ···················· | ···················· | ···················· | ···················· |
| B | 1522 hCoV-19/USA/IL-NM084/2020\|EPI_ISL_444575\|2020-03-1 | ···················· | ···················· | ···················· | ···················· | ···················· | ···················· |
| B | 1523 hCoV-19/USA/IL-NM085/2020\|EPI_ISL_444576\|2020-03-1 | ···················· | ···················· | ···················· | ···················· | ···················· | ···················· |
| B | 1524 hCoV-19/USA/IL-NM087/2020\|EPI_ISL_444577\|2020-03-1 | ···················· | ···················· | ···················· | ···················· | ···················· | ···················· |
| B | 1525 hCoV-19/Wales/PHWC-2BC38/2020\|EPI_ISL_445823\|2020- | ···················· | ···················· | ···················· | ···················· | ···················· | ···················· |
| B | 1526 hCoV-19/Wales/PHWC-2BC47/2020\|EPI_ISL_445824\|2020- | ···················· | ···················· | ···················· | ···················· | ···················· | ···················· |
| B | 1527 hCoV-19/Wales/PHWC-2BCB0/2020\|EPI_ISL_445831\|2020- | ···················· | ···················· | ···················· | ···················· | ···················· | ···················· |
| B | 1528 hCoV-19/Wales/PHWC-2BCDE/2020\|EPI_ISL_445833\|2020- | ···················· | ···················· | ···················· | ···················· | ···················· | ···················· |
| B | 1529 hCoV-19/Wales/PHWC-2BD62/2020\|EPI_ISL_445842\|2020- | ···················· | ····A··············· | ···················· | ···················· | ···················· | ···················· |
| B | 1530 hCoV-19/Wales/PHWC-2BE41/2020\|EPI_ISL_445856\|2020- | ···················· | ···················· | ···················· | ···················· | ···················· | ···················· |
| B | 1531 hCoV-19/Wales/PHWC-2BEE7/2020\|EPI_ISL_445866\|2020- | ···················· | ···················· | ···················· | ···················· | ···················· | ···················· |
| B | 1532 hCoV-19/Wales/PHWC-2BF20/2020\|EPI_ISL_445870\|2020- | ···················· | ···················· | ···················· | ···················· | ···················· | ···················· |
| B | 1533 hCoV-19/Wales/PHWC-2C5A5/2020\|EPI_ISL_445878\|2020- | ···················· | ···················· | ···················· | ···················· | ···················· | ···················· |
| B | 1534 hCoV-19/Wales/PHWC-2C61B/2020\|EPI_ISL_445885\|2020- | ···················· | ····A··············· | ···················· | ···················· | ···················· | ···················· |
| B | 1535 hCoV-19/Wales/PHWC-2C639/2020\|EPI_ISL_445887\|2020- | ···················· | ···················· | ···················· | ···················· | ···················· | ···················· |
| B | 1536 hCoV-19/Wales/PHWC-2C6B1/2020\|EPI_ISL_445895\|2020- | ···················· | ···················· | ···················· | ···················· | ···················· | ···················· |
| B | 1537 hCoV-19/Fujian/IM3520002T/2020\|EPI_ISL_431180\|2020 | ···················· | ···················· | ···················· | ···················· | ···················· | ···················· |
| B | 1538 hCoV-19/England/SHEF-C9E36/2020\|EPI_ISL_432478\|202 | ···················· | ···················· | ···················· | ···················· | ···················· | ···················· |
| B | 1539 hCoV-19/England/SHEF-CA020/2020\|EPI_ISL_432480\|202 | ···················· | ···················· | ···················· | ···················· | ···················· | ···················· |
| B | 1540 hCoV-19/England/SHEF-CA0B7/2020\|EPI_ISL_432481\|202 | ···················· | ···················· | ···················· | ···················· | ···················· | ···················· |
| B | 1541 hCoV-19/England/SHEF-CA1F0/2020\|EPI_ISL_432487\|202 | ···················· | ···················· | ···················· | ···················· | ···················· | ···················· |
| B | 1542 hCoV-19/Austria/Graz-MUG11/2020\|EPI_ISL_437300\|202 | ···················· | ···················· | ···················· | ···················· | ···················· | ···················· |
| B | 1543 hCoV-19/Austria/Graz-MUG15/2020\|EPI_ISL_437301\|202 | ···················· | ···················· | ···················· | ···················· | ···················· | ···················· |
| B | 1544 hCoV-19/Austria/Graz-MUG16/2020\|EPI_ISL_437302\|202 | ···················· | ···················· | ···················· | ···················· | ···················· | ···················· |
| B | 1545 hCoV-19/Austria/Graz-MUG17/2020\|EPI_ISL_437303\|202 | ···················· | ···················· | ···················· | ···················· | ···················· | ···················· |
| B | 1546 hCoV-19/Turkey/HSGM-674/2020\|EPI_ISL_437304\|2020-0 | ···················· | ···················· | ···················· | ···················· | ···················· | ···················· |
| B | 1547 hCoV-19/Turkey/HSGM-1418/2020\|EPI_ISL_437305\|2020- | ···················· | ···················· | ···················· | ···················· | ···················· | ···················· |
| B | 1548 hCoV-19/Turkey/HSGM-1450/2020\|EPI_ISL_437306\|2020- | ···················· | ···················· | ···················· | ···················· | ···················· | ···················· |
| B | 1549 hCoV-19/Turkey/HSGM-1081/2020\|EPI_ISL_437310\|2020- | ···················· | ·A·················· | ···················· | ···················· | ···················· | ··A················· |
| B | 1550 hCoV-19/Turkey/HSGM-1428/2020\|EPI_ISL_437311\|2020- | ···················· | ···················· | ···················· | ···················· | ···················· | ···················· |
| B | 1551 hCoV-19/Turkey/HSGM-1468/2020\|EPI_ISL_437312\|2020- | ···················· | ···················· | ···················· | ···················· | ···················· | ···················· |
| B | 1552 hCoV-19/Turkey/HSGM-302/2020\|EPI_ISL_437313\|2020-0 | ···················· | ···················· | ···················· | ···················· | ···················· | ···················· |

| | | | | | | | |
|---|---|---|---|---|---|---|---|
| B | 1553 hCoV-19/Turkey/HSGM-942/2020\|EPI_ISL_437315\|2020-0 | ···················· | ···················· | ···················· | ···················· | ···················· | ···················· |
| B | 1554 hCoV-19/Turkey/HSGM-1444/2020\|EPI_ISL_437318\|2020- | ···················· | ···················· | ···················· | ···················· | ···················· | ···················· |
| B | 1555 hCoV-19/Turkey/HSGM-1490/2020\|EPI_ISL_437319\|2020- | ···················· | ···················· | ···················· | ···················· | ···················· | ···················· |
| B | 1556 hCoV-19/Turkey/HSGM-1492/2020\|EPI_ISL_437320\|2020- | ···················· | ···················· | ···················· | ···················· | ···················· | ···················· |
| B | 1557 hCoV-19/Turkey/HSGM-1497/2020\|EPI_ISL_437321\|2020- | ···················· | ···················· | ···················· | ···················· | ···················· | ···················· |
| B | 1558 hCoV-19/Turkey/HSGM-1009/2020\|EPI_ISL_437322\|2020- | ···················· | ···················· | ···················· | ···················· | ···················· | ···················· |
| B | 1559 hCoV-19/Turkey/HSGM-1481/2020\|EPI_ISL_437323\|2020- | ···················· | ···················· | ···················· | ···················· | ···················· | ···················· |
| B | 1560 hCoV-19/Turkey/HSGM-1476/2020\|EPI_ISL_437324\|2020- | ···················· | ···················· | ···················· | ···················· | ···················· | ···················· |
| B | 1561 hCoV-19/Turkey/HSGM-1495/2020\|EPI_ISL_437325\|2020- | ···················· | ···················· | ···················· | ···················· | ···················· | ···················· |
| B | 1562 hCoV-19/Turkey/HSGM-1432/2020\|EPI_ISL_437326\|2020- | ···················· | ·A·················· | ···················· | ···················· | ···················· | ··A················· |
| B | 1563 hCoV-19/Turkey/HSGM-1458/2020\|EPI_ISL_437327\|2020- | ···················· | ···················· | ···················· | ···················· | ···················· | ···················· |
| B | 1564 hCoV-19/Turkey/HSGM-7668/2020\|EPI_ISL_437328\|2020- | ···················· | ···················· | ···················· | ···················· | ···················· | ···················· |
| B | 1565 hCoV-19/Turkey/HSGM-6204/2020\|EPI_ISL_437329\|2020- | ···················· | ···················· | ···················· | ···················· | ···················· | ···················· |
| B | 1566 hCoV-19/Turkey/HSGM-1452/2020\|EPI_ISL_437330\|2020- | ···················· | ···················· | ···················· | ···················· | ···················· | ···················· |
| B | 1567 hCoV-19/Turkey/HSGM-1014/2020\|EPI_ISL_437331\|2020- | ···················· | ···················· | ···················· | ···················· | ···················· | ···················· |
| B | 1568 hCoV-19/Turkey/HSGM-4698/2020\|EPI_ISL_437332\|2020- | ···················· | ···················· | ···················· | ···················· | ···················· | ···················· |
| B | 1569 hCoV-19/Turkey/HSGM-11640/2020\|EPI_ISL_437333\|2020 | ···················· | ···················· | ···················· | ···················· | ···················· | ···················· |
| B | 1570 hCoV-19/Turkey/HSGM-10232/2020\|EPI_ISL_437334\|2020 | ···················· | ···················· | ···················· | ···················· | ···················· | ···················· |
| B | 1571 hCoV-19/Turkey/HSGM-12059/2020\|EPI_ISL_437335\|2020 | ···················· | ···················· | ···················· | ···················· | ···················· | ···················· |
| B | 1572 hCoV-19/DRC/2496/2020\|EPI_ISL_437337\|2020-04-15 | ···················· | ···················· | ···················· | ···················· | ···················· | ···················· |
| B | 1573 hCoV-19/DRC/2529/2020\|EPI_ISL_437338\|2020-04-15 | ···················· | ···················· | ···················· | ···················· | ···················· | ···················· |
| B | 1574 hCoV-19/DRC/2536/2020\|EPI_ISL_437339\|2020-04-15 | ···················· | ···················· | ···················· | ···················· | ···················· | ···················· |
| B | 1575 hCoV-19/DRC/2563/2020\|EPI_ISL_437340\|2020-04-15 | ···················· | ···················· | ···················· | ···················· | ···················· | ···················· |
| B | 1576 hCoV-19/DRC/2580/2020\|EPI_ISL_437341\|2020-04-15 | ···················· | ···················· | ···················· | ···················· | ···················· | ···················· |
| B | 1577 hCoV-19/DRC/2670/2020\|EPI_ISL_437343\|2020-04-15 | ···················· | ···················· | ···················· | ···················· | ···················· | ···················· |
| B | 1578 hCoV-19/DRC/2813/2020\|EPI_ISL_437346\|2020-04-16 | ···················· | ···················· | ···················· | ···················· | ···················· | ···················· |
| B | 1579 hCoV-19/DRC/2827/2020\|EPI_ISL_437348\|2020-04-16 | ···················· | ···················· | ···················· | ···················· | ···················· | ···················· |
| B | 1580 hCoV-19/DRC/2855/2020\|EPI_ISL_437350\|2020-04-16 | ···················· | ···················· | ···················· | ···················· | ···················· | ···················· |
| B | 1582 hCoV-19/DRC/2904/2020\|EPI_ISL_437352\|2020-04-17 | ···················· | ···················· | ···················· | ···················· | ···················· | ···················· |
| B | 1583 hCoV-19/DRC/2939/2020\|EPI_ISL_437354\|2020-04-17 | ···················· | ···················· | ···················· | ···················· | ···················· | ···················· |
| B | 1584 hCoV-19/DRC/3041/2020\|EPI_ISL_437356\|2020-04-18 | ···················· | ···················· | ···················· | ···················· | ···················· | ···················· |
| B | 1585 hCoV-19/DRC/3070/2020\|EPI_ISL_437357\|2020-04-18 | ···················· | ···················· | ···················· | ···················· | ···················· | ···················· |
| B | 1586 hCoV-19/DRC/3089/2020\|EPI_ISL_437358\|2020-04-19 | ···················· | ···················· | ···················· | ···················· | ···················· | ···················· |
| B | 1587 hCoV-19/Germany/BY-MVP-0001/2020\|EPI_ISL_437359\|20 | ···················· | ···················· | ···················· | ···················· | ···················· | ···················· |
| B | 1588 hCoV-19/USA/MN-MDH-284/2020\|EPI_ISL_437361\|2020-03 | ···················· | ···················· | ···················· | ···················· | ···················· | ···················· |
| B | 1589 hCoV-19/USA/MN-MDH-286/2020\|EPI_ISL_437362\|2020-03 | ···················· | ···················· | ···················· | ···················· | ···················· | ···················· |
| B | 1590 hCoV-19/USA/MN-MDH-315/2020\|EPI_ISL_437363\|2020-03 | ···················· | ···················· | ···················· | ···················· | ···················· | ···················· |
| B | 1591 hCoV-19/USA/MN-MDH-336/2020\|EPI_ISL_437364\|2020-04 | ···················· | ···················· | ···················· | ···················· | ···················· | ···················· |
| B | 1592 hCoV-19/USA/MN-MDH-372/2020\|EPI_ISL_437365\|2020-04 | ···················· | ···················· | ···················· | ···················· | ···················· | ···················· |
| B | 1593 hCoV-19/USA/MN-MDH-374/2020\|EPI_ISL_437366\|2020-04 | ···················· | ···················· | ···················· | ···················· | ···················· | ···················· |
| B | 1594 hCoV-19/USA/MN-MDH-387/2020\|EPI_ISL_437367\|2020-04 | ···················· | ····G··············· | ···················· | ···················· | ···················· | ···················· |
| B | 1595 hCoV-19/USA/MN-MDH-390/2020\|EPI_ISL_437368\|2020-04 | ···················· | ···················· | ···················· | ···················· | ···················· | ···················· |
| B | 1596 hCoV-19/USA/MN-MDH-398/2020\|EPI_ISL_437369\|2020-04 | ···················· | ···················· | ···················· | ···················· | ···················· | ···················· |
| B | 1597 hCoV-19/England/CAMB-81F82/2020\|EPI_ISL_438600\|202 | ···················· | ···················· | ···················· | ···················· | ···················· | ···················· |
| B | 1598 hCoV-19/England/CAMB-81F91/2020\|EPI_ISL_438601\|202 | ···················· | ···················· | ···················· | ···················· | ···················· | ···················· |
| B | 1599 hCoV-19/England/CAMB-81FA0/2020\|EPI_ISL_438602\|202 | ···················· | ···················· | ···················· | ···················· | ···················· | ···················· |
| B | 1600 hCoV-19/England/CAMB-81FCE/2020\|EPI_ISL_438604\|202 | ···················· | ···················· | ···················· | ···················· | ···················· | ···················· |
| B | 1601 hCoV-19/England/CAMB-82006/2020\|EPI_ISL_438608\|202 | ···················· | ···················· | ···················· | ···················· | ···················· | ···················· |
| B | 1602 hCoV-19/England/CAMB-820AC/2020\|EPI_ISL_438618\|202 | ···················· | ···················· | ···················· | ···················· | ···················· | ···················· |
| B | 1603 hCoV-19/England/CAMB-820CA/2020\|EPI_ISL_438619\|202 | ···················· | ···················· | ···················· | ···················· | ···················· | ···················· |
| B | 1604 hCoV-19/England/CAMB-82103/2020\|EPI_ISL_438620\|202 | ···················· | ···················· | ···················· | ···················· | ···················· | ···················· |
| B | 1605 hCoV-19/England/CAMB-82121/2020\|EPI_ISL_438621\|202 | ···················· | ···················· | ···················· | ···················· | ···················· | ···················· |
| B | 1606 hCoV-19/England/CAMB-82130/2020\|EPI_ISL_438622\|202 | ···················· | ···················· | ···················· | ···················· | ···················· | ···················· |
| B | 1607 hCoV-19/England/CAMB-8214F/2020\|EPI_ISL_438623\|202 | ···················· | ···················· | ···················· | ···················· | ···················· | ···················· |
| B | 1608 hCoV-19/England/CAMB-8216D/2020\|EPI_ISL_438624\|202 | ···················· | ···················· | ···················· | ···················· | ···················· | ···················· |
| B | 1609 hCoV-19/England/CAMB-8217C/2020\|EPI_ISL_438625\|202 | ···················· | ···················· | ···················· | ···················· | ···················· | ···················· |
| B | 1610 hCoV-19/England/CAMB-8218B/2020\|EPI_ISL_438626\|202 | ···················· | ···················· | ···················· | ···················· | ···················· | ···················· |
| B | 1611 hCoV-19/England/CAMB-821A9/2020\|EPI_ISL_438628\|202 | ···················· | ···················· | ···················· | ···················· | ···················· | ···················· |
| B | 1612 hCoV-19/England/CAMB-821C7/2020\|EPI_ISL_438630\|202 | ···················· | ···················· | ···················· | ···················· | ···················· | ···················· |
| B | 1613 hCoV-19/England/CAMB-821E5/2020\|EPI_ISL_438631\|202 | ···················· | ···················· | ···················· | ···················· | ···················· | ···················· |
| B | 1614 hCoV-19/England/CAMB-821F4/2020\|EPI_ISL_438632\|202 | ···················· | ···················· | ···················· | ···················· | ···················· | ···················· |
| B | 1615 hCoV-19/England/CAMB-82200/2020\|EPI_ISL_438633\|202 | ···················· | ···················· | ···················· | ···················· | ···················· | ···················· |
| B | 1616 hCoV-19/England/CAMB-829DE/2020\|EPI_ISL_438644\|202 | ···················· | ···················· | ···················· | ···················· | ···················· | ···················· |
| B | 1617 hCoV-19/England/CAMB-82A44/2020\|EPI_ISL_438649\|202 | ···················· | ···················· | ···················· | ···················· | ···················· | ···················· |
| B | 1618 hCoV-19/England/CAMB-82A53/2020\|EPI_ISL_438650\|202 | ···················· | ···················· | ···················· | ···················· | ···················· | ···················· |
| B | 1619 hCoV-19/England/CAMB-82A71/2020\|EPI_ISL_438652\|202 | ···················· | ···················· | ···················· | ···················· | ···················· | ···················· |
| B | 1620 hCoV-19/England/CAMB-82ABD/2020\|EPI_ISL_438656\|202 | ···················· | ···················· | ···················· | ···················· | ···················· | ···················· |
| B | 1621 hCoV-19/England/CAMB-82ADB/2020\|EPI_ISL_438658\|202 | ···················· | ···················· | ···················· | ···················· | ···················· | ···················· |
| B | 1622 hCoV-19/England/CAMB-82B14/2020\|EPI_ISL_438660\|202 | ···················· | ···················· | ···················· | ···················· | ···················· | ···················· |
| B | 1623 hCoV-19/England/CAMB-82B32/2020\|EPI_ISL_438662\|202 | ···················· | ···················· | ···················· | ···················· | ···················· | ···················· |
| B | 1624 hCoV-19/England/CAMB-82B50/2020\|EPI_ISL_438663\|202 | ···················· | ···················· | ···················· | ···················· | ···················· | ···················· |
| B | 1625 hCoV-19/England/CAMB-82B6F/2020\|EPI_ISL_438664\|202 | ···················· | ···················· | ···················· | ···················· | ···················· | ···················· |
| B | 1626 hCoV-19/England/CAMB-82B7E/2020\|EPI_ISL_438665\|202 | ···················· | ···················· | ···················· | ···················· | ···················· | ···················· |
| B | 1627 hCoV-19/England/CAMB-82BAB/2020\|EPI_ISL_438666\|202 | ···················· | ···················· | ···················· | ···················· | ···················· | ···················· |
| B | 1628 hCoV-19/England/CAMB-82BBA/2020\|EPI_ISL_438667\|202 | ···················· | ···················· | ···················· | ···················· | ···················· | ···················· |
| B | 1629 hCoV-19/England/CAMB-82BD8/2020\|EPI_ISL_438669\|202 | ···················· | ···················· | ···················· | ···················· | ···················· | ···················· |
| B | 1630 hCoV-19/England/CAMB-82C11/2020\|EPI_ISL_438670\|202 | ···················· | ···················· | ···················· | ···················· | ···················· | ···················· |
| B | 1631 hCoV-19/England/CAMB-82C20/2020\|EPI_ISL_438671\|202 | ···················· | ···················· | ···················· | ···················· | ···················· | ···················· |

| | | | | | | | |
|---|---|---|---|---|---|---|---|
| B | 1632 hCoV-19/England/CAMB-82C3F/2020\|EPI_ISL_438672\|202 | .................... | .................... | ..................... | .................... | .................... | .................... |
| B | 1633 hCoV-19/England/CAMB-82C8A/2020\|EPI_ISL_438676\|202 | .................... | .................... | ..................... | .................... | .................... | .................... |
| B | 1634 hCoV-19/England/CAMB-82C99/2020\|EPI_ISL_438677\|202 | .................... | .................... | ..................... | .................... | .................... | .................... |
| B | 1635 hCoV-19/England/CAMB-82CA8/2020\|EPI_ISL_438678\|202 | .................... | .................... | ..................... | .................... | .................... | .................... |
| B | 1636 hCoV-19/England/CAMB-82CC6/2020\|EPI_ISL_438679\|202 | .................... | .................... | ..................... | .................... | .................... | .................... |
| B | 1637 hCoV-19/England/CAMB-83481/2020\|EPI_ISL_438681\|202 | .................... | .................... | ..................... | .................... | .................... | .................... |
| B | 1638 hCoV-19/England/CAMB-83CF2/2020\|EPI_ISL_438682\|202 | .................... | .................... | ..................... | .................... | .................... | .................... |
| B | 1639 hCoV-19/England/CAMB-83D0E/2020\|EPI_ISL_438683\|202 | .................... | .................... | ..................... | .................... | .................... | .................... |
| B | 1640 hCoV-19/England/CAMB-83D1D/2020\|EPI_ISL_438684\|202 | .................... | .................... | ..................... | .................... | .................... | .................... |
| B | 1641 hCoV-19/England/CAMB-83D59/2020\|EPI_ISL_438686\|202 | .................... | .................... | ..................... | .................... | .................... | .................... |
| B | 1642 hCoV-19/England/CAMB-83D68/2020\|EPI_ISL_438687\|202 | .................... | .................... | ..................... | .................... | .................... | .................... |
| B | 1643 hCoV-19/England/CAMB-83D77/2020\|EPI_ISL_438688\|202 | .................... | .................... | ..................... | .................... | .................... | .................... |
| B | 1644 hCoV-19/England/CAMB-83D86/2020\|EPI_ISL_438689\|202 | .................... | .................... | ..................... | .................... | .................... | .................... |
| B | 1645 hCoV-19/England/CAMB-83D95/2020\|EPI_ISL_438690\|202 | .................... | .................... | ..................... | .................... | .................... | .................... |
| B | 1646 hCoV-19/England/CAMB-83DA4/2020\|EPI_ISL_438691\|202 | .................... | .................... | ..................... | .................... | .................... | .................... |
| B | 1647 hCoV-19/England/CAMB-83DB3/2020\|EPI_ISL_438692\|202 | .................... | .................... | ..................... | .................... | .................... | .................... |
| B | 1648 hCoV-19/England/CAMB-83DC2/2020\|EPI_ISL_438693\|202 | .................... | .................... | ..................... | .................... | .................... | .................... |
| B | 1649 hCoV-19/England/CAMB-83E0B/2020\|EPI_ISL_438696\|202 | .................... | .................... | ..................... | .................... | .................... | .................... |
| B | 1650 hCoV-19/England/CAMB-83E1A/2020\|EPI_ISL_438697\|202 | .................... | .................... | ..................... | .................... | .................... | .................... |
| B | 1651 hCoV-19/England/CAMB-83E29/2020\|EPI_ISL_438698\|202 | .................... | .................... | ..................... | .................... | .................... | .................... |
| B | 1652 hCoV-19/England/CAMB-83E38/2020\|EPI_ISL_438699\|202 | .................... | .................... | ..................... | .................... | .................... | .................... |
| B | 1653 hCoV-19/England/CAMB-729D1/2020\|EPI_ISL_439900\|202 | .................... | .................... | ..................... | .................... | .................... | .................... |
| B | 1654 hCoV-19/England/CAMB-722D6/2020\|EPI_ISL_439902\|202 | .................... | .................... | ..................... | .................... | .................... | .................... |
| B | 1655 hCoV-19/England/CAMB-72652/2020\|EPI_ISL_439926\|202 | .................... | .................... | ..................... | .................... | .................... | .................... |
| B | 1656 hCoV-19/England/CAMB-71C15/2020\|EPI_ISL_439927\|202 | .................... | .................... | ..................... | .................... | .................... | .................... |
| B | 1657 hCoV-19/England/CAMB-72151/2020\|EPI_ISL_439930\|202 | .................... | .................... | ..................... | .................... | .................... | .................... |
| B | 1658 hCoV-19/England/CAMB-7286B/2020\|EPI_ISL_439931\|202 | .................... | .................... | ..................... | .................... | .................... | .................... |
| B | 1659 hCoV-19/England/CAMB-71D4F/2020\|EPI_ISL_439932\|202 | .................... | .................... | ..................... | .................... | .................... | .................... |
| B | 1660 hCoV-19/England/CAMB-71EE2/2020\|EPI_ISL_439933\|202 | .................... | .................... | ..................... | .................... | .................... | .................... |
| B | 1661 hCoV-19/England/CAMB-720DC/2020\|EPI_ISL_439934\|202 | .................... | .................... | ..................... | .................... | .................... | .................... |
| B | 1662 hCoV-19/England/CAMB-72027/2020\|EPI_ISL_439935\|202 | .................... | .................... | ..................... | .................... | .................... | .................... |
| B | 1663 hCoV-19/England/CAMB-71F67/2020\|EPI_ISL_439936\|202 | .................... | .................... | ..................... | .................... | .................... | .................... |
| B | 1664 hCoV-19/England/CAMB-72A65/2020\|EPI_ISL_439937\|202 | .................... | .................... | ..................... | .................... | .................... | .................... |
| B | 1665 hCoV-19/England/CAMB-72564/2020\|EPI_ISL_439938\|202 | .................... | .................... | ..................... | .................... | .................... | .................... |
| B | 1666 hCoV-19/England/CAMB-71CBB/2020\|EPI_ISL_439947\|202 | .................... | .................... | ..................... | .................... | .................... | .................... |
| B | 1667 hCoV-19/England/CAMB-72810/2020\|EPI_ISL_439949\|202 | .................... | .................... | ..................... | .................... | .................... | .................... |
| B | 1668 hCoV-19/England/CAMB-72F0C/2020\|EPI_ISL_439954\|202 | .................... | .................... | ..................... | .................... | .................... | .................... |
| B | 1669 hCoV-19/England/CAMB-72EF0/2020\|EPI_ISL_439956\|202 | .................... | .................... | ..................... | .................... | .................... | .................... |
| B | 1670 hCoV-19/England/BRIS-1221AA/2020\|EPI_ISL_439957\|20 | .................... | .................... | ..................... | .................... | .................... | .................... |
| B | 1671 hCoV-19/England/CAMB-72D5D/2020\|EPI_ISL_439958\|202 | .................... | .................... | ..................... | .................... | .................... | .................... |
| B | 1672 hCoV-19/England/BRIS-12230E/2020\|EPI_ISL_439959\|20 | .................... | .................... | ..................... | .................... | .................... | .................... |
| B | 1673 hCoV-19/England/CAMB-733D2/2020\|EPI_ISL_439961\|202 | .................... | .................... | ..................... | .................... | .................... | .................... |
| B | 1674 hCoV-19/England/CAMB-72C23/2020\|EPI_ISL_439963\|202 | .................... | .................... | ..................... | .................... | .................... | .................... |
| B | 1675 hCoV-19/England/BRIS-122061/2020\|EPI_ISL_439964\|20 | .................... | .................... | ..................... | .................... | .................... | .................... |
| B | 1676 hCoV-19/England/BRIS-12208F/2020\|EPI_ISL_439965\|20 | .................... | .................... | ..................... | .................... | .................... | .................... |
| B | 1677 hCoV-19/England/BRIS-121D4D/2020\|EPI_ISL_439966\|20 | .................... | .................... | ..................... | .................... | .................... | .................... |
| B | 1678 hCoV-19/England/BRIS-12217D/2020\|EPI_ISL_439967\|20 | .................... | .................... | ..................... | .................... | .................... | .................... |
| B | 1679 hCoV-19/England/CAMB-72E2D/2020\|EPI_ISL_439968\|202 | .................... | .................... | ..................... | .................... | .................... | .................... |
| B | 1680 hCoV-19/England/CAMB-73536/2020\|EPI_ISL_439970\|202 | .................... | .................... | ..................... | .................... | .................... | .................... |
| B | 1681 hCoV-19/England/BRIS-1222F2/2020\|EPI_ISL_439971\|20 | .................... | .A.................. | ..................... | .................... | .................... | .................... |
| B | 1682 hCoV-19/England/BRIS-121E0E/2020\|EPI_ISL_439972\|20 | .................... | .................... | ..................... | .................... | .................... | .................... |
| B | 1683 hCoV-19/England/BRIS-122377/2020\|EPI_ISL_439973\|20 | .................... | .................... | ..................... | .................... | .................... | .................... |
| B | 1684 hCoV-19/England/BRIS-121DF2/2020\|EPI_ISL_439974\|20 | .................... | .................... | ..................... | .................... | .................... | .................... |
| B | 1685 hCoV-19/England/CAMB-72C41/2020\|EPI_ISL_439975\|202 | .................... | .................... | ..................... | .................... | .................... | .................... |
| B | 1686 hCoV-19/England/CAMB-7334B/2020\|EPI_ISL_439980\|202 | .................... | .................... | ..................... | .................... | .................... | .................... |
| B | 1687 hCoV-19/England/BRIS-121D89/2020\|EPI_ISL_439981\|20 | .................... | .................... | ..................... | .................... | .................... | .................... |
| B | 1688 hCoV-19/England/BRIS-122070/2020\|EPI_ISL_439982\|20 | .................... | .................... | ..................... | .................... | .................... | .................... |
| B | 1689 hCoV-19/England/BRIS-121FCF/2020\|EPI_ISL_439983\|20 | .................... | .................... | ..................... | .................... | .................... | .................... |
| B | 1690 hCoV-19/England/BRIS-121D5C/2020\|EPI_ISL_439984\|20 | .................... | .................... | ..................... | .................... | .................... | .................... |
| B | 1691 hCoV-19/England/BRIS-12219B/2020\|EPI_ISL_439985\|20 | .................... | .................... | ..................... | .................... | .................... | .................... |
| B | 1692 hCoV-19/England/BRIS-121EC2/2020\|EPI_ISL_439986\|20 | .................... | .................... | ..................... | .................... | .................... | .................... |
| B | 1693 hCoV-19/England/CAMB-73651/2020\|EPI_ISL_439987\|202 | .................... | .................... | ..................... | .................... | .................... | .................... |
| B | 1694 hCoV-19/England/BRIS-121F92/2020\|EPI_ISL_439988\|20 | .................... | .................... | ..................... | .................... | .................... | .................... |
| B | 1695 hCoV-19/England/CAMB-72B71/2020\|EPI_ISL_439989\|202 | .................... | .................... | ..................... | .................... | .................... | .................... |
| B | 1696 hCoV-19/England/CAMB-72ED2/2020\|EPI_ISL_439990\|202 | .................... | .................... | ..................... | .................... | .................... | .................... |
| B | 1697 hCoV-19/England/CAMB-72CE7/2020\|EPI_ISL_439991\|202 | .................... | .................... | ..................... | .................... | .................... | .................... |
| B | 1698 hCoV-19/England/BRIS-121DC5/2020\|EPI_ISL_439992\|20 | .................... | .................... | ..................... | .................... | .................... | .................... |
| B | 1699 hCoV-19/England/CAMB-72E69/2020\|EPI_ISL_439993\|202 | .................... | .................... | ..................... | .................... | .................... | .................... |
| B | 1700 hCoV-19/England/CAMB-734B1/2020\|EPI_ISL_439997\|202 | .................... | .................... | ..................... | .................... | .................... | .................... |
| B | 1701 hCoV-19/England/BRIS-121F74/2020\|EPI_ISL_439998\|20 | .................... | .................... | ..................... | .................... | .................... | .................... |
| B | 1702 hCoV-19/England/CAMB-72DE4/2020\|EPI_ISL_439999\|202 | .................... | .................... | ..................... | .................... | .................... | .................... |
| B | 1703 hCoV-19/England/LIVE-9BCF7/2020\|EPI_ISL_440910\|202 | .................... | .................... | ..................... | .................... | .................... | .................... |
| B | 1704 hCoV-19/England/LIVE-9BD4F/2020\|EPI_ISL_440912\|202 | .................... | .................... | ..................... | .................... | .................... | .................... |
| B | 1705 hCoV-19/England/LIVE-9BDA9/2020\|EPI_ISL_440917\|202 | .................... | .................... | ..................... | .................... | .................... | .................... |
| B | 1706 hCoV-19/England/LIVE-9CA29/2020\|EPI_ISL_440930\|202 | .................... | .................... | ..................... | .................... | .................... | .................... |
| B | 1707 hCoV-19/England/LIVE-9CB44/2020\|EPI_ISL_440935\|202 | .................... | .................... | ..................... | .................... | .................... | .................... |
| B | 1708 hCoV-19/England/LIVE-9CB80/2020\|EPI_ISL_440936\|202 | .................... | .................... | ..................... | .................... | .................... | .................... |
| B | 1709 hCoV-19/England/LIVE-9CC05/2020\|EPI_ISL_440939\|202 | .................... | .................... | ..................... | .................... | .................... | .................... |

| | | | | | | | | |
|---|---|---|---|---|---|---|---|---|
| B | 1710 | hCoV-19/England/LIVE-9CE0F/2020\|EPI_ISL_440946\|202 | ···················· | ···················· | ···················· | ···················· | ···················· | ···················· |
| B | 1711 | hCoV-19/England/LIVE-9CE78/2020\|EPI_ISL_440948\|202 | ············T······· | ···················· | ···················· | ···················· | ···················· | ···················· |
| B | 1712 | hCoV-19/England/LIVE-9CE87/2020\|EPI_ISL_440949\|202 | ···················· | ···················· | ···················· | ···················· | ···················· | ···················· |
| B | 1713 | hCoV-19/England/LOND-D3516/2020\|EPI_ISL_440951\|202 | ···················· | ···················· | ···················· | ···················· | ···················· | ···················· |
| B | 1714 | hCoV-19/England/LOND-D3525/2020\|EPI_ISL_440952\|202 | ···················· | ···················· | ···················· | ···················· | ···················· | ···················· |
| B | 1715 | hCoV-19/England/LOND-D3534/2020\|EPI_ISL_440953\|202 | ···················· | ···················· | ···················· | ···················· | ···················· | ···················· |
| B | 1716 | hCoV-19/England/LOND-D3543/2020\|EPI_ISL_440954\|202 | ···················· | ···················· | ···················· | ···················· | ···················· | ···················· |
| B | 1717 | hCoV-19/England/LOND-D35BC/2020\|EPI_ISL_440960\|202 | ···················· | ···················· | ···················· | ···················· | ···················· | ···················· |
| B | 1718 | hCoV-19/England/LOND-D35E9/2020\|EPI_ISL_440962\|202 | ···················· | ···················· | ···················· | ···················· | ···················· | ···················· |
| B | 1719 | hCoV-19/England/LOND-D3604/2020\|EPI_ISL_440964\|202 | ···················· | ···················· | ···················· | ···················· | ···················· | ···················· |
| B | 1720 | hCoV-19/England/LOND-D3613/2020\|EPI_ISL_440965\|202 | ···················· | ···················· | ···················· | ···················· | ···················· | ···················· |
| B | 1721 | hCoV-19/England/LOND-D3622/2020\|EPI_ISL_440966\|202 | ···················· | ···················· | ···················· | ···················· | ···················· | ···················· |
| B | 1722 | hCoV-19/England/LOND-D3631/2020\|EPI_ISL_440967\|202 | ···················· | ···················· | ···················· | ···················· | ···················· | ···················· |
| B | 1723 | hCoV-19/England/LOND-D3640/2020\|EPI_ISL_440968\|202 | ···················· | ···················· | ···················· | ···················· | ···················· | ···················· |
| B | 1724 | hCoV-19/England/LOND-D365F/2020\|EPI_ISL_440969\|202 | ···················· | ···················· | ···················· | ···················· | ···················· | ···················· |
| B | 1725 | hCoV-19/England/LOND-D366E/2020\|EPI_ISL_440970\|202 | ···················· | ···················· | ···················· | ···················· | ···················· | ···················· |
| B | 1726 | hCoV-19/England/LOND-D368C/2020\|EPI_ISL_440971\|202 | ···················· | ···················· | ···················· | ···················· | ···················· | ···················· |
| B | 1727 | hCoV-19/England/LOND-D369B/2020\|EPI_ISL_440972\|202 | ···················· | ···················· | ···················· | ···················· | ···················· | ···················· |
| B | 1728 | hCoV-19/England/LOND-D36B9/2020\|EPI_ISL_440973\|202 | ···················· | ···················· | ···················· | ···················· | ···················· | ···················· |
| B | 1729 | hCoV-19/England/LOND-D36D7/2020\|EPI_ISL_440975\|202 | ···················· | ···················· | ···················· | ···················· | ···················· | ···················· |
| B | 1730 | hCoV-19/England/LOND-D36F5/2020\|EPI_ISL_440976\|202 | ···················· | ···················· | ···················· | ···················· | ···················· | ···················· |
| B | 1731 | hCoV-19/England/LOND-D373E/2020\|EPI_ISL_440980\|202 | ···················· | ···················· | ···················· | ···················· | ···················· | ···················· |
| B | 1732 | hCoV-19/England/LOND-D374D/2020\|EPI_ISL_440981\|202 | ···················· | ···················· | ···················· | ···················· | ···················· | ···················· |
| B | 1733 | hCoV-19/England/LOND-D3789/2020\|EPI_ISL_440983\|202 | ···················· | ···················· | ···················· | ···················· | ···················· | ···················· |
| B | 1734 | hCoV-19/England/LOND-D3798/2020\|EPI_ISL_440984\|202 | ···················· | ···················· | ···················· | ···················· | ···················· | ···················· |
| B | 1735 | hCoV-19/England/LOND-D37A7/2020\|EPI_ISL_440985\|202 | ···················· | ···················· | ···················· | ···················· | ···················· | ···················· |
| B | 1736 | hCoV-19/England/LOND-D37B6/2020\|EPI_ISL_440986\|202 | ···················· | ···················· | ···················· | ···················· | ···················· | ···················· |
| B | 1737 | hCoV-19/England/LOND-D37C5/2020\|EPI_ISL_440987\|202 | ···················· | ···················· | ···················· | ···················· | ···················· | ···················· |
| B | 1738 | hCoV-19/England/LOND-D37D4/2020\|EPI_ISL_440988\|202 | ···················· | ···················· | ···················· | ···················· | ···················· | ···················· |
| B | 1739 | hCoV-19/England/LOND-D37E3/2020\|EPI_ISL_440989\|202 | ···················· | ···················· | ···················· | ···················· | ···················· | ···················· |
| B | 1740 | hCoV-19/England/LOND-D37F2/2020\|EPI_ISL_440990\|202 | ···················· | ···················· | ···················· | ···················· | ···················· | ···················· |
| B | 1741 | hCoV-19/England/LOND-D380E/2020\|EPI_ISL_440991\|202 | ···················· | ···················· | ···················· | ···················· | ···················· | ···················· |
| B | 1742 | hCoV-19/England/LOND-D382C/2020\|EPI_ISL_440993\|202 | ···················· | ···················· | ···················· | ···················· | ···················· | ···················· |
| B | 1743 | hCoV-19/England/LOND-D383B/2020\|EPI_ISL_440994\|202 | ···················· | ···················· | ···················· | ···················· | ···················· | ···················· |
| B | 1744 | hCoV-19/England/LOND-D384A/2020\|EPI_ISL_440995\|202 | ···················· | ···················· | ···················· | ···················· | ···················· | ···················· |
| B | 1745 | hCoV-19/England/LOND-D3859/2020\|EPI_ISL_440996\|202 | ···················· | ···················· | ···················· | ···················· | ···················· | ···················· |
| B | 1746 | hCoV-19/England/LOND-D3868/2020\|EPI_ISL_440997\|202 | ···················· | ···················· | ···················· | ···················· | ···················· | ···················· |
| B | 1747 | hCoV-19/England/LOND-D3877/2020\|EPI_ISL_440998\|202 | ···················· | ···················· | ···················· | ···················· | ···················· | ···················· |
| B | 1748 | hCoV-19/England/LOND-D3886/2020\|EPI_ISL_440999\|202 | ···················· | ···················· | ···················· | ···················· | ···················· | ···················· |
| B | 1749 | hCoV-19/Singapore/98/2020\|EPI_ISL_443190\|2020-04-0 | ···················· | ···················· | ···················· | ···················· | ···················· | ···················· |
| B | 1751 | hCoV-19/Singapore/100/2020\|EPI_ISL_443192\|2020-04- | ···················· | ···················· | ···················· | ···················· | ···················· | ···················· |
| B | 1752 | hCoV-19/Singapore/101/2020\|EPI_ISL_443193\|2020-03- | ···················· | ···················· | ···················· | ···················· | ···················· | ···················· |
| B | 1753 | hCoV-19/Singapore/103/2020\|EPI_ISL_443195\|2020-03- | ···················· | ···················· | ···················· | ···················· | ···················· | ···················· |
| B | 1754 | hCoV-19/Singapore/105/2020\|EPI_ISL_443197\|2020-03- | ···················· | ···················· | ···················· | ···················· | ···················· | ···················· |
| B | 1755 | hCoV-19/Singapore/106/2020\|EPI_ISL_443198\|2020-03- | ···················· | ···················· | ···················· | ···················· | ···················· | ···················· |
| B | 1756 | hCoV-19/Singapore/107/2020\|EPI_ISL_443199\|2020-03- | ···················· | ···················· | ···················· | ···················· | ···················· | ···················· |
| B | 1757 | hCoV-19/India/GJ-GBRC26/2020\|EPI_ISL_444457\|2020-0 | ···················· | ···················· | ···················· | ···················· | ···················· | ···················· |
| B | 1758 | hCoV-19/India/GJ-GBRC27/2020\|EPI_ISL_444458\|2020-0 | ···················· | ···················· | ···················· | ···················· | ···················· | ···················· |
| B | 1759 | hCoV-19/India/GJ-GBRC28/2020\|EPI_ISL_444459\|2020-0 | ···················· | ·A·················· | ···················· | ···················· | ···················· | ···A················ |
| B | 1760 | hCoV-19/India/GJ-GBRC29/2020\|EPI_ISL_444460\|2020-0 | ···················· | ···················· | ···················· | ···················· | ···················· | ···················· |
| B | 1761 | hCoV-19/India/GJ-GBRC30/2020\|EPI_ISL_444461\|2020-0 | ···················· | ·A·················· | ···················· | ···················· | ···················· | ···A················ |
| B | 1762 | hCoV-19/India/GJ-GBRC31/2020\|EPI_ISL_444462\|2020-0 | ···················· | ···················· | ···················· | ···················· | ···················· | ···················· |
| B | 1763 | hCoV-19/India/GJ-GBRC32/2020\|EPI_ISL_444463\|2020-0 | ···················· | ···················· | ···················· | ···················· | ···················· | ···················· |
| B | 1764 | hCoV-19/India/GJ-GBRC33/2020\|EPI_ISL_444464\|2020-0 | ···T················ | ·A·················· | ···················· | ···················· | ···················· | ···A················ |
| B | 1765 | hCoV-19/India/GJ-GBRC34/2020\|EPI_ISL_444465\|2020-0 | ···················· | ·A·················· | ···················· | ···················· | ···················· | ···A················ |
| B | 1766 | hCoV-19/India/GJ-GBRC35/2020\|EPI_ISL_444466\|2020-0 | ···················· | ·A·················· | ···················· | ···················· | ···················· | ···A················ |
| B | 1767 | hCoV-19/India/GJ-GBRC36/2020\|EPI_ISL_444467\|2020-0 | ···················· | ···················· | ···················· | ···················· | ···················· | ···················· |
| B | 1768 | hCoV-19/India/GJ-GBRC37/2020\|EPI_ISL_444468\|2020-0 | ···················· | ···················· | ···················· | ···················· | ···················· | ···················· |
| B | 1769 | hCoV-19/India/GJ-GBRC38/2020\|EPI_ISL_444469\|2020-0 | ···················· | ·A·················· | ···················· | ···················· | ···················· | ···A················ |
| B | 1770 | hCoV-19/India/GJ-GBRC39/2020\|EPI_ISL_444470\|2020-0 | ···················· | ···················· | ···················· | ···················· | ···················· | ···················· |
| B | 1771 | hCoV-19/India/GJ-GBRC40/2020\|EPI_ISL_444471\|2020-0 | ···················· | ···················· | ···················· | ···················· | ···················· | ···················· |
| B | 1772 | hCoV-19/India/GJ-GBRC41/2020\|EPI_ISL_444472\|2020-0 | ···················· | ·A·················· | ···················· | ···················· | ···················· | ···A················ |
| B | 1773 | hCoV-19/India/GJ-GBRC42/2020\|EPI_ISL_444473\|2020-0 | ···················· | ···················· | ···················· | ···················· | ···················· | ···················· |
| B | 1774 | hCoV-19/India/GJ-GBRC43/2020\|EPI_ISL_444474\|2020-0 | ···················· | ···················· | ···················· | ···················· | ···················· | ···················· |
| B | 1775 | hCoV-19/India/GJ-GBRC44/2020\|EPI_ISL_444475\|2020-0 | ···················· | ···················· | ···················· | ···················· | ···················· | ···················· |
| B | 1776 | hCoV-19/India/GJ-GBRC45/2020\|EPI_ISL_444476\|2020-0 | ···················· | ·A·················· | ···················· | ···················· | ···················· | ···A················ |
| B | 1777 | hCoV-19/India/GJ-GBRC46/2020\|EPI_ISL_444477\|2020-0 | ···················· | ···················· | ···················· | ···················· | ···················· | ···················· |
| B | 1778 | hCoV-19/India/GJ-GBRC47/2020\|EPI_ISL_444478\|2020-0 | ···················· | ···················· | ···················· | ···················· | ···················· | ···················· |
| B | 1779 | hCoV-19/India/GJ-GBRC48/2020\|EPI_ISL_444479\|2020-0 | ···················· | ···················· | ···················· | ···················· | ···················· | ···················· |
| B | 1780 | hCoV-19/India/GJ-GBRC49/2020\|EPI_ISL_444480\|2020-0 | ···················· | ···················· | ···················· | ···················· | ···················· | ···················· |
| B | 1781 | hCoV-19/India/GJ-GBRC50/2020\|EPI_ISL_444481\|2020-0 | ···················· | ···················· | ···················· | ···················· | ···················· | ···················· |
| B | 1782 | hCoV-19/India/GJ-GBRC51/2020\|EPI_ISL_444482\|2020-0 | ···················· | ···················· | ···················· | ···················· | ···················· | ···················· |
| B | 1783 | hCoV-19/India/GJ-GBRC52/2020\|EPI_ISL_444483\|2020-0 | ···················· | ···················· | ···················· | ···················· | ···················· | ···················· |
| B | 1784 | hCoV-19/India/GJ-GBRC53/2020\|EPI_ISL_444484\|2020-0 | ···················· | ···················· | ···················· | ···················· | ···················· | ···················· |
| B | 1785 | hCoV-19/India/GJ-GBRC54/2020\|EPI_ISL_444485\|2020-0 | ···················· | ···················· | ···················· | ···················· | ···················· | ···················· |
| B | 1786 | hCoV-19/India/GJ-GBRC55/2020\|EPI_ISL_444486\|2020-0 | ···················· | ·A·················· | ···················· | ···················· | ···················· | ···A················ |
| B | 1787 | hCoV-19/Sweden/KI-CTMR-V300056977-1H/2020\|EPI_ISL_ | ···················· | ···················· | ···················· | ···················· | ···················· | ···················· |
| B | 1788 | hCoV-19/Sweden/KI-CTMR-V300057201-8E/2020\|EPI_ISL_ | ···················· | ···················· | ···················· | ···················· | ···················· | ···················· |

| | | | | | | | | |
|---|---|---|---|---|---|---|---|---|
| B | 1789 hCoV-19/Sweden/KI-CTMR-V300056977-4A/2020\|EPI_ISL_ | ···················· | ···················· | ···················· | ···················· | ···················· | ···················· | |
| B | 1790 hCoV-19/Uruguay/Mdeo-1/2020\|EPI_ISL_444493\|2020-03 | ···················· | ···················· | ···················· | ···················· | ···················· | ···················· | |
| B | 1791 hCoV-19/Canada/QC_AA3/2020\|EPI_ISL_444495\|2020-04- | ···················· | ···················· | ···················· | ···················· | ···················· | ···················· | |
| B | 1792 hCoV-19/Wales/PHWC-2BA10/2020\|EPI_ISL_445794\|2020- | ···················· | ····A··············· | ···················· | ···················· | ···················· | ···················· | |
| B | 1793 hCoV-19/England/SHEF-D0731/2020\|EPI_ISL_432614\|202 | ···················· | ···················· | ···················· | ···················· | ···················· | ···················· | |
| B | 1794 hCoV-19/England/SHEF-D0D99/2020\|EPI_ISL_432620\|202 | ···················· | ···················· | ···················· | ···················· | ···················· | ···················· | |
| B | 1795 hCoV-19/England/SHEF-D1053/2020\|EPI_ISL_432622\|202 | ···················· | ···················· | ···················· | ···················· | ···················· | ···················· | |
| B | 1796 hCoV-19/England/SHEF-D1536/2020\|EPI_ISL_432630\|202 | ···················· | ···················· | ···················· | ···················· | ···················· | ···················· | |
| B | 1797 hCoV-19/England/SHEF-D1E2C/2020\|EPI_ISL_432642\|202 | ···················· | ···················· | ···················· | ···················· | ···················· | ···················· | |
| B | 1798 hCoV-19/England/SHEF-D1F56/2020\|EPI_ISL_432644\|202 | ···················· | ···················· | ···················· | ···················· | ···················· | ···················· | |
| B | 1799 hCoV-19/England/NOTT-10FA3F/2020\|EPI_ISL_432675\|20 | ···················· | ···················· | ···················· | ···················· | ···················· | ···················· | |
| B | 1800 hCoV-19/England/NOTT-10FB69/2020\|EPI_ISL_432691\|20 | ···················· | ···················· | ···················· | ···················· | ···················· | ···················· | |
| B | 1801 hCoV-19/England/CAMB-7AF7D/2020\|EPI_ISL_433900\|202 | ···················· | ···················· | ···················· | ···················· | ···················· | ···················· | |
| B | 1802 hCoV-19/England/CAMB-7AFAA/2020\|EPI_ISL_433902\|202 | ···················· | ···················· | ···················· | ···················· | ···················· | ···················· | |
| B | 1803 hCoV-19/England/CAMB-7AFF5/2020\|EPI_ISL_433904\|202 | ···················· | ···················· | ···················· | ···················· | ···················· | ···················· | |
| B | 1804 hCoV-19/England/CAMB-7B000/2020\|EPI_ISL_433905\|202 | ···················· | ···················· | ···················· | ···················· | ···················· | ···················· | |
| B | 1805 hCoV-19/England/CAMB-7B01F/2020\|EPI_ISL_433906\|202 | ···················· | ···················· | ···················· | ···················· | ···················· | ···················· | |
| B | 1806 hCoV-19/England/CAMB-7B02E/2020\|EPI_ISL_433907\|202 | ···················· | ···················· | ···················· | ···················· | ···················· | ···················· | |
| B | 1807 hCoV-19/England/CAMB-7B06A/2020\|EPI_ISL_433909\|202 | ···················· | ···················· | ···················· | ···················· | ···················· | ···················· | |
| B | 1808 hCoV-19/England/CAMB-7B079/2020\|EPI_ISL_433910\|202 | ···················· | ···················· | ···················· | ···················· | ···················· | ···················· | |
| B | 1809 hCoV-19/England/CAMB-7B088/2020\|EPI_ISL_433911\|202 | ···················· | ···················· | ···················· | ···················· | ···················· | ···················· | |
| B | 1810 hCoV-19/England/CAMB-7B0A6/2020\|EPI_ISL_433913\|202 | ···················· | ···················· | ···················· | ···················· | ···················· | ···················· | |
| B | 1811 hCoV-19/England/CAMB-7B0B5/2020\|EPI_ISL_433914\|202 | ···················· | ···················· | ···················· | ···················· | ···················· | ···················· | |
| B | 1812 hCoV-19/England/CAMB-7B0C4/2020\|EPI_ISL_433915\|202 | ···················· | ···················· | ···················· | ···················· | ···················· | ···················· | |
| B | 1813 hCoV-19/England/CAMB-7B0F1/2020\|EPI_ISL_433918\|202 | ···················· | ···················· | ···················· | ···················· | ···················· | ···················· | |
| B | 1814 hCoV-19/England/CAMB-7B10D/2020\|EPI_ISL_433919\|202 | ···················· | ···················· | ···················· | ···················· | ···················· | ···················· | |
| B | 1815 hCoV-19/England/CAMB-7B11C/2020\|EPI_ISL_433920\|202 | ···················· | ···················· | ···················· | ···················· | ···················· | ···················· | |
| B | 1816 hCoV-19/England/CAMB-7B13A/2020\|EPI_ISL_433922\|202 | ···················· | ···················· | ···················· | ···················· | ···················· | ···················· | |
| B | 1817 hCoV-19/England/CAMB-7B149/2020\|EPI_ISL_433923\|202 | ···················· | ···················· | ···················· | ···················· | ···················· | ···················· | |
| B | 1818 hCoV-19/England/CAMB-7B194/2020\|EPI_ISL_433926\|202 | ···················· | ···················· | ···················· | ···················· | ···················· | ···················· | |
| B | 1819 hCoV-19/England/CAMB-7B1A3/2020\|EPI_ISL_433927\|202 | ···················· | ···················· | ···················· | ···················· | ···················· | ···················· | |
| B | 1820 hCoV-19/England/CAMB-7B1B2/2020\|EPI_ISL_433928\|202 | ···················· | ···················· | ···················· | ···················· | ···················· | ···················· | |
| B | 1821 hCoV-19/England/CAMB-7B1C1/2020\|EPI_ISL_433929\|202 | ···················· | ···················· | ···················· | ···················· | ···················· | ···················· | |
| B | 1822 hCoV-19/England/CAMB-7B1D0/2020\|EPI_ISL_433930\|202 | ···················· | ···················· | ···················· | ···················· | ···················· | ···················· | |
| B | 1823 hCoV-19/England/CAMB-7B1EF/2020\|EPI_ISL_433931\|202 | ···················· | ···················· | ···················· | ···················· | ···················· | ···················· | |
| B | 1824 hCoV-19/England/CAMB-7B237/2020\|EPI_ISL_433933\|202 | ···················· | ···················· | ···················· | ···················· | ···················· | ···················· | |
| B | 1825 hCoV-19/England/CAMB-7B273/2020\|EPI_ISL_433934\|202 | ···················· | ···················· | ···················· | ···················· | ···················· | ···················· | |
| B | 1826 hCoV-19/England/CAMB-7B291/2020\|EPI_ISL_433935\|202 | ···················· | ···················· | ···················· | ···················· | ···················· | ···················· | |
| B | 1827 hCoV-19/England/CAMB-7B2A0/2020\|EPI_ISL_433936\|202 | ···················· | ···················· | ···················· | ···················· | ···················· | ···················· | |
| B | 1828 hCoV-19/England/CAMB-7B2BF/2020\|EPI_ISL_433937\|202 | ···················· | ···················· | ···················· | ···················· | ···················· | ···················· | |
| B | 1829 hCoV-19/England/CAMB-7B2CE/2020\|EPI_ISL_433938\|202 | ···················· | ···················· | ···················· | ···················· | ···················· | ···················· | |
| B | 1830 hCoV-19/England/CAMB-7B2EC/2020\|EPI_ISL_433939\|202 | ···················· | ···················· | ···················· | ···················· | ···················· | ···················· | |
| B | 1831 hCoV-19/England/CAMB-7B325/2020\|EPI_ISL_433942\|202 | ···················· | ···················· | ···················· | ···················· | ···················· | ···················· | |
| B | 1832 hCoV-19/England/CAMB-7B343/2020\|EPI_ISL_433944\|202 | ···················· | ···················· | ···················· | ···················· | ···················· | ···················· | |
| B | 1833 hCoV-19/England/CAMB-7B361/2020\|EPI_ISL_433945\|202 | ···················· | ···················· | ···················· | ···················· | ···················· | ···················· | |
| B | 1834 hCoV-19/England/CAMB-7B39E/2020\|EPI_ISL_433948\|202 | ···················· | ···················· | ···················· | ···················· | ···················· | ···················· | |
| B | 1835 hCoV-19/England/CAMB-7B3BC/2020\|EPI_ISL_433949\|202 | ···················· | ···················· | ···················· | ···················· | ···················· | ···················· | |
| B | 1836 hCoV-19/England/CAMB-7B3CB/2020\|EPI_ISL_433950\|202 | ···················· | ···················· | ···················· | ···················· | ···················· | ···················· | |
| B | 1837 hCoV-19/England/CAMB-7B3DA/2020\|EPI_ISL_433951\|202 | ···················· | ···················· | ···················· | ···················· | ···················· | ···················· | |
| B | 1838 hCoV-19/England/CAMB-7B3E9/2020\|EPI_ISL_433952\|202 | ···················· | ···················· | ···················· | ···················· | ···················· | ···················· | |
| B | 1839 hCoV-19/England/CAMB-7B3F8/2020\|EPI_ISL_433953\|202 | ···················· | ···················· | ···················· | ···················· | ···················· | ···················· | |
| B | 1840 hCoV-19/England/CAMB-7B404/2020\|EPI_ISL_433954\|202 | ···················· | ···················· | ···················· | ···················· | ···················· | ···················· | |
| B | 1841 hCoV-19/England/CAMB-7B413/2020\|EPI_ISL_433955\|202 | ···················· | ···················· | ···················· | ···················· | ···················· | ···················· | |
| B | 1842 hCoV-19/England/CAMB-7B422/2020\|EPI_ISL_433956\|202 | ···················· | ···················· | ···················· | ···················· | ···················· | ···················· | |
| B | 1843 hCoV-19/England/CAMB-7B431/2020\|EPI_ISL_433957\|202 | ···················· | ···················· | ···················· | ···················· | ···················· | ···················· | |
| B | 1844 hCoV-19/England/CAMB-7B440/2020\|EPI_ISL_433958\|202 | ···················· | ···················· | ···················· | ···················· | ···················· | ···················· | |
| B | 1845 hCoV-19/England/CAMB-7B45F/2020\|EPI_ISL_433959\|202 | ···················· | ···················· | ···················· | ···················· | ···················· | ···················· | |
| B | 1846 hCoV-19/England/CAMB-7B48C/2020\|EPI_ISL_433961\|202 | ···················· | ···················· | ···················· | ···················· | ···················· | ···················· | |
| B | 1847 hCoV-19/England/CAMB-7B49B/2020\|EPI_ISL_433962\|202 | ···················· | ···G················ | ···················· | ···················· | ···················· | ···················· | |
| B | 1848 hCoV-19/England/CAMB-7B4AA/2020\|EPI_ISL_433963\|202 | ···················· | ···················· | ···················· | ···················· | ···················· | ···················· | |
| B | 1849 hCoV-19/England/CAMB-7B4B9/2020\|EPI_ISL_433964\|202 | ···················· | ···················· | ···················· | ···················· | ···················· | ···················· | |
| B | 1850 hCoV-19/England/CAMB-7B4C8/2020\|EPI_ISL_433965\|202 | ···················· | ···················· | ···················· | ···················· | ···················· | ···················· | |
| B | 1851 hCoV-19/England/CAMB-7B4D7/2020\|EPI_ISL_433966\|202 | ···················· | ···················· | ···················· | ···················· | ···················· | ···················· | |
| B | 1852 hCoV-19/England/CAMB-7B4E6/2020\|EPI_ISL_433967\|202 | ···················· | ···················· | ···················· | ···················· | ···················· | ···················· | |
| B | 1853 hCoV-19/England/CAMB-7B4F5/2020\|EPI_ISL_433968\|202 | ···················· | ···················A | ···················· | ···················· | ···················· | ···················· | 1 |
| B | 1854 hCoV-19/England/CAMB-7B501/2020\|EPI_ISL_433969\|202 | ···················· | ···················· | ···················· | ···················· | ···················· | ···················· | |
| B | 1855 hCoV-19/England/CAMB-7B510/2020\|EPI_ISL_433970\|202 | ···················· | ···················· | ···················· | ···················· | ···················· | ···················· | |
| B | 1856 hCoV-19/England/CAMB-7B52F/2020\|EPI_ISL_433971\|202 | ···················· | ···················· | ···················· | ···················· | ···················· | ···················· | |
| B | 1857 hCoV-19/England/CAMB-7B53E/2020\|EPI_ISL_433972\|202 | ···················· | ···················· | ···················· | ···················· | ···················· | ···················· | |
| B | 1858 hCoV-19/England/CAMB-7B54D/2020\|EPI_ISL_433973\|202 | ···················· | ···················· | ···················· | ···················· | ···················· | ···················· | |
| B | 1859 hCoV-19/England/CAMB-7B55C/2020\|EPI_ISL_433974\|202 | ···················· | ···················· | ···················· | ···················· | ···················· | ···················· | |
| B | 1860 hCoV-19/England/CAMB-7B56B/2020\|EPI_ISL_433975\|202 | ···················· | ···················· | ···················· | ···················· | ···················· | ···················· | |
| B | 1861 hCoV-19/England/CAMB-7BFD6/2020\|EPI_ISL_433976\|202 | ···················· | ···················· | ···················· | ···················· | ···················· | ···················· | |
| B | 1862 hCoV-19/England/CAMB-7C03C/2020\|EPI_ISL_433982\|202 | ···················· | ···················· | ···················· | ···················· | ···················· | ···················· | |
| B | 1863 hCoV-19/England/CAMB-7C0D2/2020\|EPI_ISL_433992\|202 | ···················· | ···················· | ···················· | ···················· | ···················· | ···················· | |
| B | 1864 hCoV-19/England/CAMB-7C148/2020\|EPI_ISL_433999\|202 | ···················· | ···················· | ···················· | ···················· | ···················· | ···················· | |
| B | 1866 hCoV-19/Saudi Arabia/KAUST-Madinah50/2020\|EPI_ISL_ | ···················· | ···················· | ···················· | ···················· | ···················· | ···················· | |
| B | 1867 hCoV-19/Saudi Arabia/KAUST-Madinah51/2020\|EPI_ISL_ | ···················· | ···················· | ···················· | ···················· | ···················· | ···················· | |

| | | | | | | | | | |
|---|---|---|---|---|---|---|---|---|---|
| B | 1868 | hCoV-19/Saudi Arabia/KAUST-Madinah53/2020\|EPI_ISL_ | .................... | .................... | .................... | .................... | .................... | .................... | |
| B | 1869 | hCoV-19/Saudi Arabia/KAUST-Madinah54/2020\|EPI_ISL_ | .................... | .................... | .................... | .................... | .................... | .................... | |
| B | 1870 | hCoV-19/Saudi Arabia/KAUST-Madinah55/2020\|EPI_ISL_ | .................... | .................... | .................... | .................... | .................... | .................... | |
| B | 1871 | hCoV-19/Saudi Arabia/KAUST-Madinah57/2020\|EPI_ISL_ | .................... | .................... | .................... | .................... | .................... | .................... | |
| B | 1872 | hCoV-19/Saudi Arabia/KAUST-Jeddah59/2020\|EPI_ISL_4 | .................... | .................... | .................... | .................... | .................... | .................... | |
| B | 1873 | hCoV-19/Saudi Arabia/KAUST-Jeddah60/2020\|EPI_ISL_4 | .................... | .................... | .................... | .................... | .................... | .................... | |
| B | 1874 | hCoV-19/England/CAMB-81FDD/2020\|EPI_ISL_438605\|202 | .................... | .................... | .................... | .................... | .................... | .................... | |
| B | 1875 | hCoV-19/England/CAMB-82024/2020\|EPI_ISL_438610\|202 | .................... | .................... | .................... | .................... | .................... | .................... | |
| B | 1876 | hCoV-19/England/CAMB-71ADF/2020\|EPI_ISL_439903\|202 | .................... | .................... | .................... | .................... | .................... | .................... | |
| B | 1877 | hCoV-19/England/CAMB-723C4/2020\|EPI_ISL_439904\|202 | .................... | .................... | .................... | .................... | .................... | .................... | |
| B | 1878 | hCoV-19/England/CAMB-725BF/2020\|EPI_ISL_439905\|202 | .................... | .................... | .................... | .................... | .................... | .................... | |
| B | 1879 | hCoV-19/England/CAMB-71E88/2020\|EPI_ISL_439907\|202 | .................... | .................... | .................... | .................... | .................... | .................... | |
| B | 1880 | hCoV-19/England/CAMB-7292C/2020\|EPI_ISL_439909\|202 | .................... | .................... | .................... | .................... | .................... | .................... | |
| B | 1881 | hCoV-19/England/CAMB-72009/2020\|EPI_ISL_439911\|202 | .................... | .................... | .................... | .................... | .................... | .................... | |
| B | 1882 | hCoV-19/England/CAMB-7236A/2020\|EPI_ISL_439914\|202 | .................... | .................... | .................... | .................... | .................... | .................... | |
| B | 1883 | hCoV-19/England/CAMB-71EA6/2020\|EPI_ISL_439917\|202 | .................... | .................... | .................... | .................... | .................... | .................... | |
| B | 1884 | hCoV-19/England/CAMB-720CD/2020\|EPI_ISL_439918\|202 | .................... | .................... | .................... | .................... | .................... | .................... | |
| B | 1885 | hCoV-19/England/CAMB-72106/2020\|EPI_ISL_439919\|202 | .................... | .................... | .................... | .................... | .................... | .................... | |
| B | 1886 | hCoV-19/England/CAMB-725DD/2020\|EPI_ISL_439920\|202 | .................... | .................... | .................... | .................... | .................... | .................... | |
| B | 1887 | hCoV-19/England/CAMB-72582/2020\|EPI_ISL_439921\|202 | .................... | .................... | .................... | .................... | .................... | .................... | |
| B | 1888 | hCoV-19/England/CAMB-71EF1/2020\|EPI_ISL_439923\|202 | .................... | .................... | .................... | .................... | .................... | .................... | |
| B | 1889 | hCoV-19/England/CAMB-728B6/2020\|EPI_ISL_439941\|202 | .................... | .................... | .................... | .................... | .................... | .................... | |
| B | 1890 | hCoV-19/England/CAMB-724B2/2020\|EPI_ISL_439943\|202 | .................... | .................... | .................... | .................... | .................... | .................... | |
| B | 1891 | hCoV-19/England/CAMB-72573/2020\|EPI_ISL_439945\|202 | .................... | .................... | .................... | .................... | .................... | .................... | |
| B | 1892 | hCoV-19/England/LIVE-9BB90/2020\|EPI_ISL_440900\|202 | .................... | A................... | .................... | .................... | .................... | .................... | |
| B | 1893 | hCoV-19/England/LIVE-9BBFA/2020\|EPI_ISL_440903\|202 | .................... | ...................A | .................... | .................... | .................... | .................... | 1 |
| B | 1894 | hCoV-19/England/LIVE-9BC06/2020\|EPI_ISL_440904\|202 | .................... | .................... | .................... | .................... | .................... | .................... | |
| B | 1895 | hCoV-19/England/LIVE-9BC33/2020\|EPI_ISL_440905\|202 | .................... | .................... | .................... | .................... | .................... | .................... | |
| B | 1896 | hCoV-19/England/LIVE-9BE3D/2020\|EPI_ISL_440922\|202 | .................... | .................... | .................... | .................... | .................... | .................... | |
| B | 1897 | hCoV-19/England/LIVE-9C93B/2020\|EPI_ISL_440923\|202 | .................... | .................... | .................... | .................... | .................... | .................... | |
| B | 1898 | hCoV-19/England/LIVE-9C959/2020\|EPI_ISL_440924\|202 | .................... | .................... | .................... | .................... | .................... | .................... | |
| B | 1899 | hCoV-19/England/LIVE-9C986/2020\|EPI_ISL_440925\|202 | .................... | .................... | .................... | .................... | .................... | .................... | |
| B | 1900 | hCoV-19/England/LIVE-9C9B3/2020\|EPI_ISL_440927\|202 | .................... | .................... | .................... | .................... | .................... | .................... | |
| B | 1901 | hCoV-19/England/SHEF-C6C7B/2020\|EPI_ISL_442001\|202 | .................... | .................... | .................... | .................... | .................... | .................... | |
| B | 1902 | hCoV-19/England/SHEF-C7991/2020\|EPI_ISL_442013\|202 | .................... | .................... | .................... | .................... | .................... | .................... | |
| B | 1903 | hCoV-19/England/SHEF-C87A5/2020\|EPI_ISL_442032\|202 | .................... | .................... | .................... | .................... | .................... | .................... | |
| B | 1904 | hCoV-19/USA/MN-UMGC-1/2020\|EPI_ISL_443183\|2020-04- | .................... | .................... | .................... | .................... | .................... | .................... | |
| B | 1905 | hCoV-19/USA/WA-UW-4296/2020\|EPI_ISL_443184\|2020-03 | .................... | .................... | .................... | .................... | .................... | .................... | |
| B | 1906 | hCoV-19/USA/OR-UW-4409/2020\|EPI_ISL_443185\|2020-03 | .................... | .................... | .................... | .................... | .................... | .................... | |
| B | 1907 | hCoV-19/USA/WA-UW-4414/2020\|EPI_ISL_443186\|2020-03 | .................... | .................... | .................... | .................... | .................... | .................... | |
| B | 1908 | hCoV-19/Brunei/1/2020\|EPI_ISL_443187\|2020-03-11 | .................... | .................... | .................... | .................... | .................... | .................... | |
| B | 1909 | hCoV-19/Singapore/96/2020\|EPI_ISL_443188\|2020-03-1 | .................... | .................... | .................... | .................... | .................... | .................... | |
| B | 1910 | hCoV-19/Singapore/97/2020\|EPI_ISL_443189\|2020-03-1 | .................... | .................... | .................... | .................... | .................... | .................... | |
| B | 1911 | hCoV-19/France/IDF-3081/2020\|EPI_ISL_443300\|2020-0 | .................... | .................... | .................... | .................... | .................... | .................... | |
| B | 1912 | hCoV-19/France/IDF-3359/2020\|EPI_ISL_443301\|2020-0 | .................... | .................... | .................... | .................... | .................... | .................... | |
| B | 1913 | hCoV-19/France/IDF-3368/2020\|EPI_ISL_443302\|2020-0 | .................... | .................... | .................... | .................... | .................... | .................... | |
| B | 1914 | hCoV-19/France/IDF-5577/2020\|EPI_ISL_443303\|2020-0 | .................... | .................... | .................... | .................... | .................... | .................... | |
| B | 1915 | hCoV-19/France/IDF-5986/2020\|EPI_ISL_443305\|2020-0 | .................... | .................... | .................... | .................... | .................... | .................... | |
| B | 1916 | hCoV-19/France/IDF-6037/2020\|EPI_ISL_443306\|2020-0 | .................... | .................... | .................... | .................... | .................... | .................... | |
| B | 1917 | hCoV-19/France/BFC-6053/2020\|EPI_ISL_443307\|2020-0 | .................... | .................... | .................... | .................... | .................... | .................... | |
| B | 1918 | hCoV-19/France/GES-6058/2020\|EPI_ISL_443308\|2020-0 | .................... | .................... | .................... | .................... | .................... | .................... | |
| B | 1919 | hCoV-19/France/HDF-3669/2020\|EPI_ISL_443309\|2020-0 | .................... | .................... | .................... | .................... | .................... | .................... | |
| B | 1920 | hCoV-19/France/GES-4420/2020\|EPI_ISL_443310\|2020-0 | .................... | .................... | .................... | .................... | .................... | .................... | |
| B | 1921 | hCoV-19/France/IDF-4423/2020\|EPI_ISL_443311\|2020-0 | .................... | .................... | .................... | .................... | .................... | .................... | |
| B | 1922 | hCoV-19/France/CVL-4430/2020\|EPI_ISL_443312\|2020-0 | .................... | .................... | .................... | .................... | .................... | .................... | |
| B | 1923 | hCoV-19/France/BRE-6041/2020\|EPI_ISL_443313\|2020-0 | .................... | .................... | .................... | .................... | .................... | .................... | |
| B | 1924 | hCoV-19/France/IDF-6073/2020\|EPI_ISL_443314\|2020-0 | .................... | .................... | .................... | .................... | .................... | .................... | |
| B | 1925 | hCoV-19/France/HDF-6466/2020\|EPI_ISL_443315\|2020-0 | .................... | .................... | .................... | .................... | .................... | .................... | |
| B | 1926 | hCoV-19/France/HDF-3668/2020\|EPI_ISL_443316\|2020-0 | .................... | .................... | .................... | .................... | .................... | .................... | |
| B | 1927 | hCoV-19/France/IDF-4432/2020\|EPI_ISL_443317\|2020-0 | .................... | .................... | .................... | .................... | .................... | .................... | |
| B | 1928 | hCoV-19/England/CAMB-83B8C/2020\|EPI_ISL_443319\|202 | .................... | .................... | .................... | .................... | .................... | .................... | |
| B | 1929 | hCoV-19/England/CAMB-83B22/2020\|EPI_ISL_443332\|202 | .................... | .................... | .................... | .................... | .................... | .................... | |
| B | 1930 | hCoV-19/England/CAMB-82F81/2020\|EPI_ISL_443335\|202 | .................... | .................... | .................... | .................... | .................... | .................... | |
| B | 1931 | hCoV-19/England/CAMB-83B6E/2020\|EPI_ISL_443336\|202 | .................... | .................... | .................... | .................... | .................... | .................... | |
| B | 1932 | hCoV-19/England/CAMB-83AAD/2020\|EPI_ISL_443338\|202 | .................... | .................... | .................... | .................... | .................... | .................... | |
| B | 1933 | hCoV-19/England/CAMB-83ADA/2020\|EPI_ISL_443340\|202 | .................... | .................... | .................... | .................... | .................... | .................... | |
| B | 1934 | hCoV-19/England/CAMB-83AF8/2020\|EPI_ISL_443342\|202 | .................... | .................... | .................... | .................... | .................... | .................... | |
| B | 1935 | hCoV-19/England/CAMB-83BE6/2020\|EPI_ISL_443343\|202 | .................... | .................... | .................... | .................... | .................... | .................... | |
| B | 1936 | hCoV-19/England/CAMB-83BAA/2020\|EPI_ISL_443345\|202 | .................... | .................... | .................... | .................... | .................... | .................... | |
| B | 1937 | hCoV-19/England/CAMB-83BC8/2020\|EPI_ISL_443349\|202 | .................... | .................... | .................... | .................... | .................... | .................... | |
| B | 1938 | hCoV-19/England/CAMB-82E75/2020\|EPI_ISL_443353\|202 | .................... | .................... | .................... | .................... | .................... | .................... | |
| B | 1939 | hCoV-19/England/CAMB-83982/2020\|EPI_ISL_443359\|202 | .................... | .................... | .................... | .................... | .................... | .................... | |
| B | 1940 | hCoV-19/England/CAMB-83BF5/2020\|EPI_ISL_443360\|202 | .................... | .................... | .................... | .................... | .................... | .................... | |
| B | 1941 | hCoV-19/USA/IL-NM0117/2020\|EPI_ISL_444600\|2020-03- | .................... | .................... | .................... | .................... | .................... | .................... | |
| B | 1942 | hCoV-19/USA/IL-NM0118/2020\|EPI_ISL_444601\|2020-03- | .................... | .................... | .................... | .................... | .................... | .................... | |
| B | 1943 | hCoV-19/USA/IL-NM0120/2020\|EPI_ISL_444603\|2020-03- | .................... | .................... | .................... | .................... | .................... | .................... | |
| B | 1944 | hCoV-19/USA/IL-NM0121/2020\|EPI_ISL_444604\|2020-03- | .................... | .................... | .................... | .................... | .................... | .................... | |
| B | 1945 | hCoV-19/USA/IL-NM0122/2020\|EPI_ISL_444605\|2020-03- | .................... | .................... | .................... | .................... | .................... | .................... | |

| | | | | | | | | |
|---|---|---|---|---|---|---|---|---|
| B | 1946 | hCoV-19/USA/IL-NM0124/2020\|EPI_ISL_444606\|2020-03- | ···················· | ···················· | ···················· | ···················· | ···················· | ···················· |
| B | 1947 | hCoV-19/USA/IL-NM0125/2020\|EPI_ISL_444607\|2020-03- | ···················· | ···················· | ···················· | ···················· | ···················· | ···················· |
| B | 1948 | hCoV-19/USA/IL-NM0126/2020\|EPI_ISL_444608\|2020-03- | ···················· | ···················· | ···················· | ···················· | ···················· | ···················· |
| B | 1949 | hCoV-19/USA/IL-NM0127/2020\|EPI_ISL_444609\|2020-03- | ···················· | ···················· | ···················· | ···················· | ···················· | ···················· |
| B | 1950 | hCoV-19/Guam/GU-NHG-02/2020\|EPI_ISL_444610\|2020-03 | ···················· | ···················· | ···················· | ···················· | ···················· | ···················· |
| B | 1951 | hCoV-19/Australia/QLDID937/2020\|EPI_ISL_444611\|202 | ···················· | ···················· | ···················· | ···················· | ···················· | ···················· |
| B | 1952 | hCoV-19/Australia/QLDID938/2020\|EPI_ISL_444612\|202 | ···················· | ···················· | ···················· | ···················· | ···················· | ···················· |
| B | 1953 | hCoV-19/USA/NY-NYUMC496/2020\|EPI_ISL_444613\|2020-0 | ···················· | ···················· | ···················· | ···················· | ···················· | ···················· |
| B | 1954 | hCoV-19/USA/NY-NYUMC498/2020\|EPI_ISL_444615\|2020-0 | ···················· | ···················· | ···················· | ···················· | ···················· | ···················· |
| B | 1955 | hCoV-19/USA/NY-NYUMC499/2020\|EPI_ISL_444616\|2020-0 | ···················· | ···················· | ···················· | ···················· | ···················· | ···················· |
| B | 1956 | hCoV-19/USA/NY-NYUMC500/2020\|EPI_ISL_444617\|2020-0 | ···················· | ···················· | ···················· | ···················· | ···················· | ···················· |
| B | 1957 | hCoV-19/USA/NY-NYUMC501/2020\|EPI_ISL_444618\|2020-0 | ···················· | ···················· | ···················· | ···················· | ···················· | ···················· |
| B | 1958 | hCoV-19/USA/NY-NYUMC502/2020\|EPI_ISL_444619\|2020-0 | ···················· | ···················· | ···················· | ···················· | ···················· | ···················· |
| B | 1959 | hCoV-19/USA/NY-NYUMC503/2020\|EPI_ISL_444620\|2020-0 | ···················· | ···················· | ···················· | ···················· | ···················· | ···················· |
| B | 1960 | hCoV-19/USA/NY-NYUMC504/2020\|EPI_ISL_444621\|2020-0 | ···················· | ···················· | ···················· | ···················· | ···················· | ···················· |
| B | 1961 | hCoV-19/USA/NY-NYUMC505/2020\|EPI_ISL_444622\|2020-0 | ···················· | ···················· | ···················· | ···················· | ···················· | ···················· |
| B | 1962 | hCoV-19/USA/NY-NYUMC506/2020\|EPI_ISL_444623\|2020-0 | ···················· | ···················· | ···················· | ···················· | ···················· | ···················· |
| B | 1963 | hCoV-19/USA/NY-NYUMC507/2020\|EPI_ISL_444624\|2020-0 | ···················· | ···················· | ···················· | ···················· | ···················· | ···················· |
| B | 1964 | hCoV-19/USA/NY-NYUMC508/2020\|EPI_ISL_444625\|2020-0 | ···················· | ···················· | ···················· | ···················· | ···················· | ···················· |
| B | 1965 | hCoV-19/USA/NY-NYUMC509/2020\|EPI_ISL_444626\|2020-0 | ···················· | ···················· | ···················· | ···················· | ···················· | ···················· |
| B | 1966 | hCoV-19/USA/NY-NYUMC510/2020\|EPI_ISL_444627\|2020-0 | ···················· | ···················· | ···················· | ···················· | ···················· | ···················· |
| B | 1967 | hCoV-19/USA/NY-NYUMC511/2020\|EPI_ISL_444628\|2020-0 | ···················· | ···················· | ···················· | ···················· | ···················· | ···················· |
| B | 1968 | hCoV-19/USA/NY-NYUMC512/2020\|EPI_ISL_444629\|2020-0 | ···················· | ···················· | ···················· | ···················· | ···················· | ···················· |
| B | 1969 | hCoV-19/USA/NY-NYUMC513/2020\|EPI_ISL_444630\|2020-0 | ···················· | ···················· | ···················· | ···················· | ···················· | ···················· |
| B | 1970 | hCoV-19/USA/NY-NYUMC514/2020\|EPI_ISL_444631\|2020-0 | ···················· | ···················· | ···················· | ···················· | ···················· | ···················· |
| B | 1971 | hCoV-19/USA/NY-NYUMC515/2020\|EPI_ISL_444632\|2020-0 | ···················· | ···················· | ···················· | ···················· | ···················· | ···················· |
| B | 1972 | hCoV-19/USA/NY-NYUMC516/2020\|EPI_ISL_444633\|2020-0 | ···················· | ···················· | ···················· | ···················· | ···················· | ···················· |
| B | 1973 | hCoV-19/USA/NY-NYUMC517/2020\|EPI_ISL_444634\|2020-0 | ···················· | ···················· | ···················· | ···················· | ···················· | ···················· |
| B | 1974 | hCoV-19/USA/NY-NYUMC518/2020\|EPI_ISL_444635\|2020-0 | ···················· | ··············C····· | ···················· | ···················· | ···················· | ···················· |
| B | 1975 | hCoV-19/USA/NY-NYUMC519/2020\|EPI_ISL_444636\|2020-0 | ···················· | ···················· | ···················· | ···················· | ···················· | ···················· |
| B | 1976 | hCoV-19/USA/NY-NYUMC520/2020\|EPI_ISL_444637\|2020-0 | ···················· | ···················· | ···················· | ···················· | ···················· | ···················· |
| B | 1977 | hCoV-19/USA/NY-NYUMC521/2020\|EPI_ISL_444638\|2020-0 | ···················· | ···················· | ···················· | ···················· | ···················· | ···················· |
| B | 1978 | hCoV-19/USA/NY-NYUMC522/2020\|EPI_ISL_444639\|2020-0 | ···················· | ···················· | ···················· | ···················· | ···················· | ···················· |
| B | 1979 | hCoV-19/USA/NY-NYUMC523/2020\|EPI_ISL_444640\|2020-0 | ···················· | ···················· | ···················· | ···················· | ···················· | ···················· |
| B | 1980 | hCoV-19/USA/NY-NYUMC524/2020\|EPI_ISL_444641\|2020-0 | ···················· | ···················· | ···················· | ···················· | ···················· | ···················· |
| B | 1981 | hCoV-19/USA/NY-NYUMC525/2020\|EPI_ISL_444642\|2020-0 | ···················· | ···················· | ···················· | ···················· | ···················· | ···················· |
| B | 1982 | hCoV-19/USA/NY-NYUMC526/2020\|EPI_ISL_444643\|2020-0 | ···················· | ···················· | ···················· | ···················· | ···················· | ···················· |
| B | 1983 | hCoV-19/USA/NY-NYUMC527/2020\|EPI_ISL_444644\|2020-0 | ···················· | ···················· | ···················· | ···················· | ···················· | ···················· |
| B | 1984 | hCoV-19/USA/NY-NYUMC528/2020\|EPI_ISL_444645\|2020-0 | ···················· | ···················· | ···················· | ···················· | ···················· | ···················· |
| B | 1985 | hCoV-19/USA/NY-NYUMC529/2020\|EPI_ISL_444646\|2020-0 | ···················· | ···················· | ···················· | ···················· | ···················· | ···················· |
| B | 1986 | hCoV-19/USA/NY-NYUMC530/2020\|EPI_ISL_444647\|2020-0 | ···················· | ···················· | ···················· | ···················· | ···················· | ···················· |
| B | 1987 | hCoV-19/USA/NY-NYUMC531/2020\|EPI_ISL_444648\|2020-0 | ···················· | ···················· | ···················· | ···················· | ···················· | ···················· |
| B | 1988 | hCoV-19/USA/NY-NYUMC533/2020\|EPI_ISL_444650\|2020-0 | ···················· | ···················· | ···················· | ···················· | ···················· | ···················· |
| B | 1989 | hCoV-19/USA/NY-NYUMC534/2020\|EPI_ISL_444651\|2020-0 | ···················· | ···················· | ···················· | ···················· | ···················· | ···················· |
| B | 1990 | hCoV-19/USA/NY-NYUMC535/2020\|EPI_ISL_444652\|2020-0 | ···················· | ···················· | ···················· | ···················· | ···················· | ···················· |
| B | 1991 | hCoV-19/USA/NY-NYUMC536/2020\|EPI_ISL_444653\|2020-0 | ···················· | ···················· | ···················· | ···················· | ···················· | ···················· |
| B | 1992 | hCoV-19/USA/NY-NYUMC537/2020\|EPI_ISL_444654\|2020-0 | ···················· | ···················· | ···················· | ···················· | ···················· | ···················· |
| B | 1993 | hCoV-19/USA/NJ-NYUMC538/2020\|EPI_ISL_444655\|2020-0 | ···················· | ···················· | ···················· | ···················· | ···················· | ···················· |
| B | 1994 | hCoV-19/USA/NY-NYUMC539/2020\|EPI_ISL_444656\|2020-0 | ···················· | ···················· | ···················· | ···················· | ···················· | ···················· |
| B | 1995 | hCoV-19/USA/NY-NYUMC540/2020\|EPI_ISL_444657\|2020-0 | ···················· | ···················· | ···················· | ···················· | ···················· | ···················· |
| B | 1996 | hCoV-19/USA/NY-NYUMC541/2020\|EPI_ISL_444658\|2020-0 | ···················· | ···················· | ···················· | ···················· | ···················· | ···················· |
| B | 1997 | hCoV-19/USA/NY-NYUMC542/2020\|EPI_ISL_444659\|2020-0 | ···················· | ···················· | ···················· | ···················· | ···················· | ···················· |
| B | 1998 | hCoV-19/USA/NY-NYUMC543/2020\|EPI_ISL_444660\|2020-0 | ···················· | ···················· | ···················· | ···················· | ···················· | ···················· |
| B | 1999 | hCoV-19/USA/NY-NYUMC544/2020\|EPI_ISL_444661\|2020-0 | ···················· | ···················· | ···················· | ···················· | ···················· | ···················· |
| B | 2000 | hCoV-19/USA/NY-NYUMC545/2020\|EPI_ISL_444662\|2020-0 | ···················· | ···················· | ···················· | ···················· | ···················· | ···················· |
| B | 2001 | hCoV-19/USA/NY-NYUMC546/2020\|EPI_ISL_444663\|2020-0 | ···················· | ···················· | ···················· | ···················· | ···················· | ···················· |
| B | 2002 | hCoV-19/USA/NY-NYUMC547/2020\|EPI_ISL_444664\|2020-0 | ···················· | ···················· | ···················· | ···················· | ···················· | ···················· |
| B | 2003 | hCoV-19/USA/NY-NYUMC548/2020\|EPI_ISL_444665\|2020-0 | ···················· | ···················· | ···················· | ···················· | ···················· | ···················· |
| B | 2004 | hCoV-19/USA/NY-NYUMC549/2020\|EPI_ISL_444666\|2020-0 | ···················· | ···················· | ···················· | ···················· | ···················· | ···················· |
| B | 2005 | hCoV-19/USA/NY-NYUMC550/2020\|EPI_ISL_444667\|2020-0 | ···················· | ···················· | ···················· | ···················· | ···················· | ···················· |
| B | 2006 | hCoV-19/USA/NY-NYUMC551/2020\|EPI_ISL_444668\|2020-0 | ···················· | ···················· | ···················· | ···················· | ···················· | ···················· |
| B | 2007 | hCoV-19/USA/NY-NYUMC552/2020\|EPI_ISL_444669\|2020-0 | ···················· | ···················· | ···················· | ···················· | ···················· | ···················· |
| B | 2008 | hCoV-19/USA/NJ-NYUMC553/2020\|EPI_ISL_444670\|2020-0 | ···················· | ···················· | ···················· | ···················· | ···················· | ···················· |
| B | 2009 | hCoV-19/USA/NY-NYUMC554/2020\|EPI_ISL_444671\|2020-0 | ···················· | ···················· | ···················· | ···················· | ···················· | ···················· |
| B | 2010 | hCoV-19/USA/NY-NYUMC556/2020\|EPI_ISL_444673\|2020-0 | ···················· | ···················· | ···················· | ···················· | ···················· | ···················· |
| B | 2011 | hCoV-19/USA/NY-NYUMC558/2020\|EPI_ISL_444675\|2020-0 | ···················· | ···················· | ···················· | ···················· | ···················· | ···················· |
| B | 2012 | hCoV-19/USA/NY-NYUMC560/2020\|EPI_ISL_444677\|2020-0 | ···················· | ···················· | ···················· | ···················· | ···················· | ···················· |
| B | 2013 | hCoV-19/USA/NY-NYUMC561/2020\|EPI_ISL_444678\|2020-0 | ···················· | ···················· | ···················· | ···················· | ···················· | ···················· |
| B | 2014 | hCoV-19/USA/NY-NYUMC562/2020\|EPI_ISL_444679\|2020-0 | ···················· | ···················· | ···················· | ···················· | ···················· | ···················· |
| B | 2015 | hCoV-19/USA/NY-NYUMC563/2020\|EPI_ISL_444680\|2020-0 | ···················· | ···················· | ···················· | ···················· | ···················· | ···················· |
| B | 2016 | hCoV-19/USA/NY-NYUMC564/2020\|EPI_ISL_444681\|2020-0 | ···················· | ···················· | ···················· | ···················· | ···················· | ···················· |
| B | 2017 | hCoV-19/USA/NY-NYUMC565/2020\|EPI_ISL_444682\|2020-0 | ···················· | ···················· | ···················· | ···················· | ···················· | ···················· |
| B | 2018 | hCoV-19/USA/NY-NYUMC566/2020\|EPI_ISL_444683\|2020-0 | ···················· | ···················· | ···················· | ···················· | ···················· | ···················· |
| B | 2019 | hCoV-19/USA/NY-NYUMC567/2020\|EPI_ISL_444684\|2020-0 | ···················· | ···················· | ···················· | ···················· | ···················· | ···················· |
| B | 2020 | hCoV-19/USA/NY-NYUMC568/2020\|EPI_ISL_444685\|2020-0 | ···················· | ···················· | ···················· | ···················· | ···················· | ···················· |
| B | 2021 | hCoV-19/USA/NY-NYUMC569/2020\|EPI_ISL_444686\|2020-0 | ···················· | ···················· | ···················· | ···················· | ···················· | ···················· |
| B | 2022 | hCoV-19/USA/NY-NYUMC570/2020\|EPI_ISL_444687\|2020-0 | ···················· | ···················· | ···················· | ···················· | ···················· | ···················· |
| B | 2023 | hCoV-19/USA/NY-NYUMC571/2020\|EPI_ISL_444688\|2020-0 | ···················· | ···················· | ···················· | ···················· | ···················· | ···················· |

| | | | | | | | |
|---|---|---|---|---|---|---|---|
| B | 2024 hCoV-19/USA/NY-NYUMC573/2020\|EPI_ISL_444690\|2020-0 | ···················· | ···················· | ····················· | ···················· | ···················· | ···················· |
| B | 2025 hCoV-19/USA/NY-NYUMC574/2020\|EPI_ISL_444691\|2020-0 | ···················· | ···················· | ····················· | ···················· | ···················· | ···················· |
| B | 2026 hCoV-19/USA/NY-NYUMC575/2020\|EPI_ISL_444692\|2020-0 | ···················· | ···················· | ····················· | ···················· | ···················· | ···················· |
| B | 2027 hCoV-19/USA/NY-NYUMC576/2020\|EPI_ISL_444693\|2020-0 | ···················· | ···················· | ····················· | ···················· | ···················· | ···················· |
| B | 2028 hCoV-19/USA/NY-NYUMC577/2020\|EPI_ISL_444694\|2020-0 | ···················· | ···················· | ····················· | ···················· | ···················· | ···················· |
| B | 2029 hCoV-19/USA/NY-NYUMC578/2020\|EPI_ISL_444695\|2020-0 | ···················· | ···················· | ····················· | ···················· | ···················· | ···················· |
| B | 2030 hCoV-19/USA/NY-NYUMC579/2020\|EPI_ISL_444696\|2020-0 | ···················· | ···················· | ····················· | ···················· | ···················· | ···················· |
| B | 2031 hCoV-19/USA/NY-NYUMC580/2020\|EPI_ISL_444697\|2020-0 | ···················· | ···················· | ····················· | ···················· | ···················· | ···················· |
| B | 2032 hCoV-19/USA/NY-NYUMC581/2020\|EPI_ISL_444698\|2020-0 | ···················· | ···················· | ····················· | ···················· | ···················· | ···················· |
| B | 2033 hCoV-19/Wales/PHWC-2C9C7/2020\|EPI_ISL_445931\|2020- | ···················· | ···················· | ····················· | ···················· | ···················· | ···················· |
| B | 2034 hCoV-19/Wales/PHWC-2CA4C/2020\|EPI_ISL_445938\|2020- | ···················· | ····A··············· | ····················· | ···················· | ···················· | ···················· |
| B | 2035 hCoV-19/Wales/PHWC-2CAA6/2020\|EPI_ISL_445944\|2020- | ···················· | ···················· | ····················· | ···················· | ···················· | ···················· |
| B | 2036 hCoV-19/Wales/PHWC-2CAB5/2020\|EPI_ISL_445945\|2020- | ···················· | ···················· | ····················· | ···················· | ···················· | ···················· |
| B | 2037 hCoV-19/Wales/PHWC-2CAC4/2020\|EPI_ISL_445946\|2020- | ···················· | ···················· | ····················· | ···················· | ···················· | ···················· |
| B | 2038 hCoV-19/Wales/PHWC-2CB1C/2020\|EPI_ISL_445950\|2020- | ···················· | ···················· | ····················· | ···················· | ···················· | ···················· |
| B | 2039 hCoV-19/Wales/PHWC-2CB2B/2020\|EPI_ISL_445951\|2020- | ···················· | ···················· | ····················· | ···················· | ···················· | ···················· |
| B | 2040 hCoV-19/Wales/PHWC-2D3B9/2020\|EPI_ISL_445980\|2020- | ···················· | ···················· | ····················· | ···················· | ···················· | ···················· |
| B | 2041 hCoV-19/Wuhan/WHUH005/2020\|EPI_ISL_449480\|2020-01- | ···················· | ···················· | ····················· | ···················· | ···················· | ···················· |
| B | 2042 hCoV-19/Wuhan/WHUH006/2020\|EPI_ISL_449481\|2020-01- | ···················· | ···················· | ····················· | ···················· | ···················· | ···················· |
| B | 2043 hCoV-19/Wuhan/WHUH011/2020\|EPI_ISL_449484\|2020-01- | ···················· | ···················· | ····················· | ···················· | ···················· | ···················· |
| B | 2044 hCoV-19/Wuhan/WHUH020/2020\|EPI_ISL_449486\|2020-01- | ···················· | ···················· | ····················· | ···················· | ···················· | ···················· |
| B | 2045 hCoV-19/USA/CA-CZB-59X002/2020\|EPI_ISL_450484\|2020 | ···················· | ···················· | ····················· | ···················· | ···················· | ···················· |
| B | 2046 hCoV-19/USA/CA-CZB-59X001/2020\|EPI_ISL_450485\|2020 | ···················· | ···················· | ····················· | ···················· | ···················· | ···················· |
| B | 2047 hCoV-19/USA/CA-CZB-59X004/2020\|EPI_ISL_450486\|2020 | ···················· | ···················· | ····················· | ···················· | ···················· | ···················· |
| B | 2048 hCoV-19/USA/CA-CZB-59X003/2020\|EPI_ISL_450487\|2020 | ···················· | ···················· | ····················· | ···················· | ···················· | ···················· |
| B | 2050 hCoV-19/Australia/VIC1157/2020\|EPI_ISL_430598\|2020 | ···················· | ···················· | ····················· | ···················· | ···················· | ···················· |
| B | 2051 hCoV-19/England/SHEF-CFE03/2020\|EPI_ISL_432601\|202 | ···················· | ···················· | ····················· | ···················· | ···················· | ···················· |
| B | 2052 hCoV-19/England/SHEF-CFE21/2020\|EPI_ISL_432602\|202 | ···················· | ···················· | ····················· | ···················· | ···················· | ···················· |
| B | 2053 hCoV-19/England/CAMB-7B307/2020\|EPI_ISL_433940\|202 | ···················· | ···················· | ····················· | ···················· | ···················· | ···················· |
| B | 2054 hCoV-19/Taiwan/TSGH-16/2020\|EPI_ISL_436101\|2020-03 | ···················· | ···················· | ····················· | ···················· | ···················· | ···················· |
| B | 2055 hCoV-19/Taiwan/TSGH-17/2020\|EPI_ISL_436102\|2020-03 | ···················· | ···················· | ····················· | ···················· | ···················· | ···················· |
| B | 2056 hCoV-19/Taiwan/TSGH-19/2020\|EPI_ISL_436104\|2020-03 | ···················· | ···················· | ····················· | ···················· | ···················· | ···················· |
| B | 2057 hCoV-19/Taiwan/TSGH-21/2020\|EPI_ISL_436106\|2020-03 | ···················· | ···················· | ····················· | ···················· | ···················· | ···················· |
| B | 2058 hCoV-19/Taiwan/TSGH-22/2020\|EPI_ISL_436107\|2020-03 | ···················· | ···················· | ····················· | ···················· | ···················· | ···················· |
| B | 2059 hCoV-19/Taiwan/TSGH-23/2020\|EPI_ISL_436108\|2020-04 | ···················· | ···················· | ····················· | ···················· | ···················· | ···················· |
| B | 2066 hCoV-19/India/KA-nimh-1598/2020\|EPI_ISL_436157\|202 | ···················· | ···················· | ····················· | ···················· | ···················· | ···················· |
| B | 2067 hCoV-19/USA/VA-DCLS-0184/2020\|EPI_ISL_437400\|2020- | ···················· | ···················· | ····················· | ···················· | ···················· | ···················· |
| B | 2068 hCoV-19/USA/VA-DCLS-0185/2020\|EPI_ISL_437401\|2020- | ···················· | ···················· | ····················· | ···················· | ···················· | ···················· |
| B | 2069 hCoV-19/USA/VA-DCLS-0187/2020\|EPI_ISL_437403\|2020- | ···················· | ···················· | ····················· | ···················· | ···················· | ···················· |
| B | 2070 hCoV-19/USA/VA-DCLS-0188/2020\|EPI_ISL_437404\|2020- | ···················· | ···················· | ····················· | ···················· | ···················· | ···················· |
| B | 2071 hCoV-19/USA/VA-DCLS-0189/2020\|EPI_ISL_437405\|2020- | ···················· | ···················· | ····················· | ···················· | ···················· | ···················· |
| B | 2072 hCoV-19/USA/VA-DCLS-0190/2020\|EPI_ISL_437406\|2020- | ···················· | ···················· | ····················· | ···················· | ···················· | ···················· |
| B | 2073 hCoV-19/USA/VA-DCLS-0191/2020\|EPI_ISL_437407\|2020- | ···················· | ···················· | ····················· | ···················· | ···················· | ···················· |
| B | 2074 hCoV-19/USA/VA-DCLS-0193/2020\|EPI_ISL_437408\|2020- | ···················· | ···················· | ····················· | ···················· | ···················· | ···················· |
| B | 2075 hCoV-19/USA/VA-DCLS-0194/2020\|EPI_ISL_437409\|2020- | ···················· | ···················· | ····················· | ···················· | ···················· | ···················· |
| B | 2076 hCoV-19/USA/VA-DCLS-0195/2020\|EPI_ISL_437410\|2020- | ···················· | ···················· | ····················· | ···················· | ···················· | ···················· |
| B | 2077 hCoV-19/USA/VA-DCLS-0196/2020\|EPI_ISL_437411\|2020- | ···················· | ···················· | ····················· | ···················· | ···················· | ···················· |
| B | 2078 hCoV-19/USA/VA-DCLS-0197/2020\|EPI_ISL_437412\|2020- | ···················· | ···················· | ····················· | ···················· | ···················· | ···················· |
| B | 2079 hCoV-19/USA/VA-DCLS-0198/2020\|EPI_ISL_437413\|2020- | ···················· | ···················· | ····················· | ···················· | ···················· | ···················· |
| B | 2080 hCoV-19/USA/VA-DCLS-0199/2020\|EPI_ISL_437414\|2020- | ···················· | ···················· | ····················· | ···················· | ···················· | ···················· |
| B | 2081 hCoV-19/USA/VA-DCLS-0200/2020\|EPI_ISL_437415\|2020- | ···················· | ···················· | ····················· | ···················· | ···················· | ···················· |
| B | 2082 hCoV-19/USA/VA-DCLS-0201/2020\|EPI_ISL_437416\|2020- | ···················· | ···················· | ····················· | ···················· | ···················· | ···················· |
| B | 2083 hCoV-19/USA/VA-DCLS-0202/2020\|EPI_ISL_437417\|2020- | ···················· | ···················· | ····················· | ···················· | ···················· | ···················· |
| B | 2084 hCoV-19/USA/VA-DCLS-0203/2020\|EPI_ISL_437418\|2020- | ···················· | ···················· | ····················· | ···················· | ···················· | ···················· |
| B | 2085 hCoV-19/USA/VA-DCLS-0204/2020\|EPI_ISL_437419\|2020- | ···················· | ···················· | ····················· | ···················· | ···················· | ···················· |
| B | 2086 hCoV-19/USA/VA-DCLS-0205/2020\|EPI_ISL_437420\|2020- | ···················· | ···················· | ····················· | ···················· | ···················· | ···················· |
| B | 2087 hCoV-19/USA/VA-DCLS-0206/2020\|EPI_ISL_437421\|2020- | ···················· | ···················· | ····················· | ···················· | ···················· | ···················· |
| B | 2088 hCoV-19/USA/VA-DCLS-0207/2020\|EPI_ISL_437422\|2020- | ···················· | ···················· | ····················· | ···················· | ···················· | ···················· |
| B | 2089 hCoV-19/USA/VA-DCLS-0208/2020\|EPI_ISL_437423\|2020- | ···················· | ···················· | ····················· | ···················· | ···················· | ···················· |
| B | 2090 hCoV-19/USA/VA-DCLS-0209/2020\|EPI_ISL_437424\|2020- | ···················· | ···················· | ····················· | ···················· | ···················· | ···················· |
| B | 2091 hCoV-19/USA/VA-DCLS-0210/2020\|EPI_ISL_437425\|2020- | ···················· | ···················· | ····················· | ···················· | ···················· | ···················· |
| B | 2092 hCoV-19/USA/VA-DCLS-0211/2020\|EPI_ISL_437426\|2020- | ···················· | ···················· | ····················· | ···················· | ···················· | ···················· |
| B | 2093 hCoV-19/USA/VA-DCLS-0212/2020\|EPI_ISL_437427\|2020- | ···················· | ···················· | ····················· | ···················· | ···················· | ···················· |
| B | 2094 hCoV-19/USA/VA-DCLS-0213/2020\|EPI_ISL_437428\|2020- | ···················· | ···················· | ····················· | ···················· | ···················· | ···················· |
| B | 2095 hCoV-19/USA/VA-DCLS-0214/2020\|EPI_ISL_437429\|2020- | ···················· | ···················· | ····················· | ···················· | ···················· | ···················· |
| B | 2096 hCoV-19/USA/VA-DCLS-0216/2020\|EPI_ISL_437431\|2020- | ···················· | ···················· | ····················· | ···················· | ···················· | ···················· |
| B | 2097 hCoV-19/USA/VA-DCLS-0217/2020\|EPI_ISL_437432\|2020- | ···················· | ···················· | ····················· | ···················· | ···················· | ···················· |
| B | 2099 hCoV-19/Serbia/KV26-04/2020\|EPI_ISL_437435\|2020-04 | ···················· | ···················· | ····················· | ···················· | ···················· | ···················· |
| B | 2100 hCoV-19/Serbia/NP363-04/2020\|EPI_ISL_437436\|2020-0 | ···················· | ···················· | ····················· | ···················· | ···················· | ···················· |
| B | 2101 hCoV-19/USA/AK-PHL200/2020\|EPI_ISL_437437\|2020-04- | ···················· | ···················· | ····················· | ···················· | ···················· | ···················· |
| B | 2102 hCoV-19/India/GJ-GBRC10/2020\|EPI_ISL_437438\|2020-0 | ···················· | ···················· | ····················· | ···················· | ···················· | ···················· |
| B | 2103 hCoV-19/India/GJ-GBRC12/2020\|EPI_ISL_437440\|2020-0 | ···················· | ···················· | ····················· | ···················· | ···················· | ···················· |
| B | 2104 hCoV-19/India/GJ-GBRC13/2020\|EPI_ISL_437441\|2020-0 | ···················· | ···················· | ····················· | ···················· | ···················· | ···················· |
| B | 2105 hCoV-19/India/GJ-GBRC14/2020\|EPI_ISL_437442\|2020-0 | ···················· | ···················· | ····················· | ···················· | ···················· | ···················· |
| B | 2106 hCoV-19/India/GJ-GBRC16/2020\|EPI_ISL_437444\|2020-0 | ···················· | ···················· | ····················· | ···················· | ···················· | ···················· |
| B | 2107 hCoV-19/India/GJ-GBRC17a/2020\|EPI_ISL_437445\|2020- | ···················· | ·A·················· | ····················· | ···················· | ···················· | ···A················ |
| B | 2108 hCoV-19/India/GJ-GBRC17b/2020\|EPI_ISL_437446\|2020- | ···················· | ·A·················· | ····················· | ···················· | ···················· | ···A················ |
| B | 2109 hCoV-19/India/GJ-GBRC18/2020\|EPI_ISL_437447\|2020-0 | ···················· | ···················· | ····················· | ···················· | ···················· | ···················· |

| | | | | | | | | |
|---|---|---|---|---|---|---|---|---|
| B | 2110 | hCoV-19/India/GJ-GBRC19/2020\|EPI_ISL_437448\|2020-0 | ···················· | ···················· | ····················· | ···················· | ···················· | ···················· |
| B | 2111 | hCoV-19/India/GJ-GBRC20/2020\|EPI_ISL_437449\|2020-0 | ···················· | ···················· | ····················· | ···················· | ···················· | ···················· |
| B | 2112 | hCoV-19/India/GJ-GBRC21/2020\|EPI_ISL_437450\|2020-0 | ···················· | ···················· | ····················· | ···················· | ···················· | ···················· |
| B | 2113 | hCoV-19/India/GJ-GBRC22/2020\|EPI_ISL_437451\|2020-0 | ···················· | ···················· | ····················· | ···················· | ···················· | ···················· |
| B | 2114 | hCoV-19/India/GJ-GBRC23/2020\|EPI_ISL_437452\|2020-0 | ···················· | ·A·················· | ····················· | ···················· | ···················· | ··A················· |
| B | 2115 | hCoV-19/India/GJ-GBRC24a/2020\|EPI_ISL_437453\|2020- | ···················· | ···················· | ····················· | ···················· | ···················· | ···················· |
| B | 2116 | hCoV-19/India/GJ-GBRC24b/2020\|EPI_ISL_437454\|2020- | ···················· | ···················· | ····················· | ···················· | ···················· | ··A················· |
| B | 2117 | hCoV-19/USA/IN-Lilly-IPB0170-14bc04/2020\|EPI_ISL_4 | ···················· | ···················· | ····················· | ···················· | ···················· | ···················· |
| B | 2118 | hCoV-19/USA/IN-Lilly-IPB0170-15bc11/2020\|EPI_ISL_4 | ···················· | ···················· | ····················· | ···················· | ···················· | ···················· |
| B | 2119 | hCoV-19/USA/IN-Lilly-IPB0170-16bc18/2020\|EPI_ISL_4 | ···················· | ···················· | ····················· | ···················· | ···················· | ···················· |
| B | 2120 | hCoV-19/USA/IN-Lilly-IPB0170-15bc12/2020\|EPI_ISL_4 | ···················· | ···A················ | ····················· | ···················· | ···················· | ···················· |
| B | 2121 | hCoV-19/Saudi Arabia/KAUST-Makkah03/2020\|EPI_ISL_4 | ···················· | ···················· | ····················· | ···················· | ···················· | ···················· |
| B | 2122 | hCoV-19/Saudi Arabia/KAUST-Makkah04/2020\|EPI_ISL_4 | ···················· | ···················· | ····················· | ···················· | ···················· | ···················· |
| B | 2123 | hCoV-19/Saudi Arabia/KAUST-Makkah05/2020\|EPI_ISL_4 | ···················· | ···················· | ····················· | ···················· | ···················· | ···················· |
| B | 2124 | hCoV-19/Saudi Arabia/KAUST-Makkah06/2020\|EPI_ISL_4 | ···················· | ···················· | ····················· | ···················· | ···················· | ···················· |
| B | 2125 | hCoV-19/Saudi Arabia/KAUST-Makkah09/2020\|EPI_ISL_4 | ···················· | ···················· | ····················· | ···················· | ···················· | ···················· |
| B | 2126 | hCoV-19/Saudi Arabia/KAUST-Makkah10/2020\|EPI_ISL_4 | ···················· | ···················· | ····················· | ···················· | ···················· | ···················· |
| B | 2127 | hCoV-19/Saudi Arabia/KAUST-Makkah12/2020\|EPI_ISL_4 | ···················· | ···················· | ····················· | ···················· | ···················· | ···················· |
| B | 2128 | hCoV-19/Saudi Arabia/KAUST-Makkah13/2020\|EPI_ISL_4 | ···················· | ···················· | ····················· | ···················· | ···················· | ···················· |
| B | 2129 | hCoV-19/Saudi Arabia/KAUST-Makkah20/2020\|EPI_ISL_4 | ···················· | ···················· | ····················· | ···················· | ···················· | ···················· |
| B | 2130 | hCoV-19/Saudi Arabia/KAUST-Madinah25/2020\|EPI_ISL_ | ···················· | ···················· | ····················· | ···················· | ···················· | ···················· |
| B | 2131 | hCoV-19/Saudi Arabia/KAUST-Madinah34/2020\|EPI_ISL_ | ···················· | ···················· | ····················· | ···················· | ···················· | ···················· |
| B | 2132 | hCoV-19/Saudi Arabia/KAUST-Madinah38/2020\|EPI_ISL_ | ···················· | ···················· | ····················· | ···················· | ···················· | ···················· |
| B | 2133 | hCoV-19/Saudi Arabia/KAUST-Madinah41/2020\|EPI_ISL_ | ···················· | ···················· | ····················· | ···················· | ···················· | ···················· |
| B | 2134 | hCoV-19/Saudi Arabia/KAUST-Madinah44/2020\|EPI_ISL_ | ···················· | ···················· | ····················· | ···················· | ···················· | ···················· |
| B | 2135 | hCoV-19/Saudi Arabia/KAUST-Madinah45/2020\|EPI_ISL_ | ···················· | ···················· | ····················· | ···················· | ···················· | ···················· |
| B | 2136 | hCoV-19/Saudi Arabia/KAUST-Jeddah64/2020\|EPI_ISL_4 | ···················· | ···················· | ····················· | ···················· | ···················· | ···················· |
| B | 2137 | hCoV-19/Saudi Arabia/KAUST-Makkah159/2020\|EPI_ISL_ | ···················· | ···················· | ····················· | ···················· | ···················· | ···················· |
| B | 2138 | hCoV-19/Saudi Arabia/KAUST-Makkah168/2020\|EPI_ISL_ | ···················· | ···················· | ····················· | ···················· | ···················· | ···················· |
| B | 2139 | hCoV-19/Saudi Arabia/KAUST-Makkah19/2020\|EPI_ISL_4 | ···················· | ···················· | ····················· | ···················· | ···················· | ···················· |
| B | 2140 | hCoV-19/Saudi Arabia/KAUST-Madinah23/2020\|EPI_ISL_ | ···················· | ···················· | ····················· | ···················· | ···················· | ···················· |
| B | 2141 | hCoV-19/Saudi Arabia/KAUST-Madinah24/2020\|EPI_ISL_ | ···················· | ···················· | ····················· | ···················· | ···················· | ···················· |
| B | 2142 | hCoV-19/Saudi Arabia/KAUST-Madinah27/2020\|EPI_ISL_ | ···················· | ···················· | ····················· | ···················· | ···················· | ···················· |
| B | 2143 | hCoV-19/Saudi Arabia/KAUST-Madinah30/2020\|EPI_ISL_ | ···················· | ···················· | ····················· | ···················· | ···················· | ···················· |
| B | 2144 | hCoV-19/Saudi Arabia/KAUST-Madinah31/2020\|EPI_ISL_ | ···················· | ···················· | ····················· | ···················· | ···················· | ···················· |
| B | 2145 | hCoV-19/Saudi Arabia/KAUST-Madinah47/2020\|EPI_ISL_ | ···················· | ···················· | ····················· | ···················· | ···················· | ···················· |
| B | 2146 | hCoV-19/Saudi Arabia/KAUST-Madinah48/2020\|EPI_ISL_ | ···················· | ···················· | ····················· | ···················· | ···················· | ···················· |
| B | 2147 | hCoV-19/Saudi Arabia/KAUST-Madinah49/2020\|EPI_ISL_ | ···················· | ···················· | ····················· | ···················· | ···················· | ···················· |
| B | 2148 | hCoV-19/England/CAMB-83E47/2020\|EPI_ISL_438700\|202 | ···················· | ···················· | ····················· | ···················· | ···················· | ···················· |
| B | 2149 | hCoV-19/England/CAMB-83E56/2020\|EPI_ISL_438701\|202 | ···················· | ···················· | ····················· | ···················· | ···················· | ···················· |
| B | 2150 | hCoV-19/England/CAMB-83E74/2020\|EPI_ISL_438702\|202 | ···················· | ···················· | ····················· | ···················· | ···················· | ···················· |
| B | 2151 | hCoV-19/England/CAMB-83E83/2020\|EPI_ISL_438703\|202 | ···················· | ···················· | ····················· | ···················· | ···················· | ···················· |
| B | 2152 | hCoV-19/England/CAMB-83EA1/2020\|EPI_ISL_438704\|202 | ···················· | ···················· | ····················· | ···················· | ···················· | ···················· |
| B | 2153 | hCoV-19/England/CAMB-83EB0/2020\|EPI_ISL_438705\|202 | ···················· | ···················· | ····················· | ···················· | ···················· | ···················· |
| B | 2154 | hCoV-19/England/CAMB-83EDE/2020\|EPI_ISL_438707\|202 | ···················· | ···················· | ····················· | ···················· | ······T············· | ···················· |
| B | 2155 | hCoV-19/England/CAMB-83EED/2020\|EPI_ISL_438708\|202 | ···················· | ···················· | ····················· | ···················· | ···················· | ···················· |
| B | 2156 | hCoV-19/England/CAMB-83EFC/2020\|EPI_ISL_438709\|202 | ···················· | ···················· | ····················· | ···················· | ···················· | ···················· |
| B | 2157 | hCoV-19/England/CAMB-83F17/2020\|EPI_ISL_438710\|202 | ···················· | ···················· | ····················· | ···················· | ···················· | ···················· |
| B | 2158 | hCoV-19/England/CAMB-83F35/2020\|EPI_ISL_438711\|202 | ···················· | ···················· | ····················· | ···················· | ···················· | ···················· |
| B | 2159 | hCoV-19/England/CAMB-83F44/2020\|EPI_ISL_438712\|202 | ···················· | ···················· | ····················· | ···················· | ···················· | ···················· |
| B | 2160 | hCoV-19/England/CAMB-83F53/2020\|EPI_ISL_438713\|202 | ···················· | ···················· | ····················· | ···················· | ···················· | ···················· |
| B | 2161 | hCoV-19/England/CAMB-83F62/2020\|EPI_ISL_438714\|202 | ···················· | ···················· | ····················· | ···················· | ···················· | ···················· |
| B | 2162 | hCoV-19/England/CAMB-83F71/2020\|EPI_ISL_438715\|202 | ···················· | ···················· | ····················· | ···················· | ···················· | ···················· |
| B | 2163 | hCoV-19/England/CAMB-83F80/2020\|EPI_ISL_438716\|202 | ···················· | ···················· | ····················· | ···················· | ···················· | ···················· |
| B | 2164 | hCoV-19/England/CAMB-83FAE/2020\|EPI_ISL_438717\|202 | ···················· | ···················· | ····················· | ···················· | ···················· | ···················· |
| B | 2165 | hCoV-19/England/CAMB-83FCC/2020\|EPI_ISL_438719\|202 | ···················· | ···················· | ····················· | ···················· | ···················· | ···················· |
| B | 2166 | hCoV-19/England/CAMB-83FDB/2020\|EPI_ISL_438720\|202 | ···················· | ···················· | ····················· | ···················· | ···················· | ···················· |
| B | 2167 | hCoV-19/England/CAMB-83FEA/2020\|EPI_ISL_438721\|202 | ···T················ | ···················· | ····················· | ···················· | ···················· | ···················· |
| B | 2168 | hCoV-19/England/CAMB-83FF9/2020\|EPI_ISL_438722\|202 | ···················· | ···················· | ····················· | ···················· | ···················· | ···················· |
| B | 2169 | hCoV-19/England/CAMB-84022/2020\|EPI_ISL_438725\|202 | ···················· | ···················· | ····················· | ···················· | ···················· | ···················· |
| B | 2170 | hCoV-19/England/CAMB-84031/2020\|EPI_ISL_438726\|202 | ···················· | ···················· | ····················· | ···················· | ···················· | ···················· |
| B | 2171 | hCoV-19/England/CAMB-8406E/2020\|EPI_ISL_438729\|202 | ···················· | ···················· | ····················· | ···················· | ···················· | ···················· |
| B | 2172 | hCoV-19/England/CAMB-840B9/2020\|EPI_ISL_438733\|202 | ···················· | ···················· | ····················· | ···················· | ···················· | ···················· |
| B | 2173 | hCoV-19/England/CAMB-840D7/2020\|EPI_ISL_438735\|202 | ···················· | ···················· | ····················· | ···················· | ···················· | ···················· |
| B | 2174 | hCoV-19/England/CAMB-840E6/2020\|EPI_ISL_438736\|202 | ···················· | ···················· | ····················· | ···················· | ···················· | ···················· |
| B | 2175 | hCoV-19/England/CAMB-840F5/2020\|EPI_ISL_438737\|202 | ···················· | ···················· | ····················· | ···················· | ···················· | ···················· |
| B | 2176 | hCoV-19/England/CAMB-8413E/2020\|EPI_ISL_438740\|202 | ···················· | ···················· | ····················· | ···················· | ···················· | ···················· |
| B | 2177 | hCoV-19/England/CAMB-8464E/2020\|EPI_ISL_438744\|202 | ···················· | ···················· | ····················· | ···················· | ···················· | ···················· |
| B | 2178 | hCoV-19/England/CAMB-8465D/2020\|EPI_ISL_438745\|202 | ···················· | ···················· | ····················· | ···················· | ···················· | ···················· |
| B | 2179 | hCoV-19/England/CAMB-8466C/2020\|EPI_ISL_438746\|202 | ···················· | ···················· | ····················· | ···················· | ···················· | ···················· |
| B | 2180 | hCoV-19/England/CAMB-8467B/2020\|EPI_ISL_438747\|202 | ···················· | ···················· | ····················· | ···················· | ···················· | ···················· |
| B | 2181 | hCoV-19/England/CAMB-8468A/2020\|EPI_ISL_438748\|202 | ···················· | ···················· | ····················· | ···················· | ···················· | ···················· |
| B | 2182 | hCoV-19/Scotland/CVR1003/2020\|EPI_ISL_438749\|2020- | ···················· | ···················· | ····················· | ···················· | ···················· | ···················· |
| B | 2183 | hCoV-19/Scotland/CVR1030/2020\|EPI_ISL_438754\|2020- | ···················· | ···················· | ····················· | ···················· | ···················· | ···················· |
| B | 2184 | hCoV-19/Scotland/CVR1043/2020\|EPI_ISL_438757\|2020- | ···················· | ···················· | ····················· | ···················· | ···················· | ···················· |
| B | 2185 | hCoV-19/Scotland/CVR1059/2020\|EPI_ISL_438760\|2020- | ···················· | ···················· | ····················· | ···················· | ···················· | ···················· |
| B | 2186 | hCoV-19/Scotland/CVR1076/2020\|EPI_ISL_438762\|2020- | ···················· | ···················· | ····················· | ···················· | ···················· | ···················· |
| B | 2187 | hCoV-19/Scotland/CVR1079/2020\|EPI_ISL_438763\|2020- | ···················· | ·A·················· | ····················· | ···················· | ···················· | ···················· |

| | | | | | | | | |
|---|---|---|---|---|---|---|---|---|
| B | 2188 | hCoV-19/Scotland/CVR1086/2020\|EPI_ISL_438764\|2020- | ···················· | ···················· | ····················· | ···················· | ···················· | ···················· |
| B | 2189 | hCoV-19/Scotland/CVR1113/2020\|EPI_ISL_438766\|2020- | ···················· | ···················· | ····················· | ···················· | ···················· | ···················· |
| B | 2190 | hCoV-19/Scotland/CVR1217/2020\|EPI_ISL_438775\|2020- | ···················· | ···················· | ····················· | ···················· | ···················· | ···················· |
| B | 2191 | hCoV-19/Scotland/CVR1221/2020\|EPI_ISL_438776\|2020- | ···················· | ···················· | ····················· | ···················· | ···················· | ···················· |
| B | 2192 | hCoV-19/Scotland/CVR1249/2020\|EPI_ISL_438779\|2020- | ···················· | ···················· | ····················· | ···················· | ···················· | ···················· |
| B | 2193 | hCoV-19/Scotland/CVR1263/2020\|EPI_ISL_438783\|2020- | ···················· | ···················· | ····················· | ···················· | ···················· | ···················· |
| B | 2194 | hCoV-19/Scotland/CVR1268/2020\|EPI_ISL_438784\|2020- | ···················· | ···················· | ····················· | ···················· | ···················· | ···················· |
| B | 2195 | hCoV-19/Scotland/CVR1402/2020\|EPI_ISL_438798\|2020- | ···················· | ···················· | ····················· | ···················· | ···················· | ···················· |
| B | 2196 | hCoV-19/England/CAMB-7A0D4/2020\|EPI_ISL_441245\|202 | ···················· | ···················· | ····················· | ···················· | ···················· | ···················· |
| B | 2197 | hCoV-19/England/CAMB-79F32/2020\|EPI_ISL_441247\|202 | ···················· | ···················· | ····················· | ···················· | ···················· | ···················· |
| B | 2198 | hCoV-19/England/CAMB-7A818/2020\|EPI_ISL_441249\|202 | ···················· | ···················· | ····················· | ···················· | ···················· | ···················· |
| B | 2199 | hCoV-19/England/CAMB-79E35/2020\|EPI_ISL_441250\|202 | ···················· | ···················· | ····················· | ···················· | ···················· | ···················· |
| B | 2200 | hCoV-19/England/CAMB-7A56C/2020\|EPI_ISL_441251\|202 | ···················· | ···················· | ····················· | ···················· | ···················· | ···················· |
| B | 2201 | hCoV-19/England/CAMB-7A48D/2020\|EPI_ISL_441253\|202 | ···················· | ···················· | ····················· | ···················· | ···················· | ···················· |
| B | 2202 | hCoV-19/England/CAMB-79BF2/2020\|EPI_ISL_441254\|202 | ···················· | ···················· | ····················· | ···················· | ···················· | ···················· |
| B | 2203 | hCoV-19/England/CAMB-7A7A2/2020\|EPI_ISL_441255\|202 | ···················· | ···················· | ····················· | ···················· | ···················· | ···················· |
| B | 2204 | hCoV-19/England/CAMB-7A678/2020\|EPI_ISL_441256\|202 | ···················· | ···················· | ····················· | ···················· | ···················· | ···················· |
| B | 2205 | hCoV-19/England/CAMB-79F23/2020\|EPI_ISL_441259\|202 | ···················· | ···················· | ····················· | ···················· | ···················· | ···················· |
| B | 2206 | hCoV-19/England/CAMB-78BA8/2020\|EPI_ISL_441260\|202 | ···················· | ···················· | ····················· | ···················· | ···················· | ···················· |
| B | 2207 | hCoV-19/England/CAMB-78E90/2020\|EPI_ISL_441262\|202 | ···················· | ···················· | ····················· | ···················· | ···················· | ···················· |
| B | 2208 | hCoV-19/England/CAMB-79F50/2020\|EPI_ISL_441266\|202 | ···················· | ···················· | ····················· | ···················· | ···················· | ···················· |
| B | 2209 | hCoV-19/England/CAMB-7E98A/2020\|EPI_ISL_442560\|202 | ···················· | ···················· | ····················· | ···················· | ···················· | ···················· |
| B | 2210 | hCoV-19/England/CAMB-816BA/2020\|EPI_ISL_442577\|202 | ···················· | ···················· | ····················· | ···················· | ···················· | ···················· |
| B | 2211 | hCoV-19/England/CAMB-816AB/2020\|EPI_ISL_442578\|202 | ···················· | ···················· | ····················· | ···················· | ···················· | ···················· |
| B | 2212 | hCoV-19/England/CAMB-814CF/2020\|EPI_ISL_442581\|202 | ···················· | ···················· | ····················· | ···················· | ···················· | ···················· |
| B | 2213 | hCoV-19/England/CAMB-7EA87/2020\|EPI_ISL_442582\|202 | ···················· | ···················· | ····················· | ···················· | ···················· | ···················· |
| B | 2214 | hCoV-19/England/CAMB-81429/2020\|EPI_ISL_442583\|202 | ···················· | ···················· | ····················· | ···················· | ···················· | ···················· |
| B | 2215 | hCoV-19/England/CAMB-7E89C/2020\|EPI_ISL_442584\|202 | ···················· | ···················· | ····················· | ···················· | ···················· | ···················· |
| B | 2216 | hCoV-19/England/CAMB-808F1/2020\|EPI_ISL_442586\|202 | ···················· | ···················· | ····················· | ···················· | ···················· | ···················· |
| B | 2217 | hCoV-19/England/CAMB-81ABE/2020\|EPI_ISL_442587\|202 | ···················· | ···················· | ····················· | ···················· | ···················· | ···················· |
| B | 2218 | hCoV-19/England/CAMB-81957/2020\|EPI_ISL_442588\|202 | ···················· | ···················· | ····················· | ···················· | ···················· | ···················· |
| B | 2219 | hCoV-19/England/CAMB-80703/2020\|EPI_ISL_442589\|202 | ···················· | ···················· | ····················· | ···················· | ···················· | ···················· |
| B | 2220 | hCoV-19/England/CAMB-7E8F6/2020\|EPI_ISL_442592\|202 | ···················· | ···················· | ····················· | ···················· | ···················· | ···················· |
| B | 2221 | hCoV-19/England/CAMB-81438/2020\|EPI_ISL_442593\|202 | ···················· | ···················· | ····················· | ···················· | ···················· | ···················· |
| B | 2222 | hCoV-19/England/CAMB-813C2/2020\|EPI_ISL_442594\|202 | ···················· | ···················· | ····················· | ···················· | ···················· | ···················· |
| B | 2223 | hCoV-19/England/CAMB-817F3/2020\|EPI_ISL_442595\|202 | ···················· | ···················· | ····················· | ···················· | ···················· | ···················· |
| B | 2224 | hCoV-19/England/CAMB-817A8/2020\|EPI_ISL_442596\|202 | ···················· | ···················· | ····················· | ···················· | ···················· | ···················· |
| B | 2225 | hCoV-19/England/CAMB-81377/2020\|EPI_ISL_442597\|202 | ···················· | ···················· | ····················· | ···················· | ···················· | ···················· |
| B | 2226 | hCoV-19/Singapore/155/2020\|EPI_ISL_443247\|2020-04- | ···················· | ···················· | ····················· | ···················· | ···················· | ···················· |
| B | 2227 | hCoV-19/Singapore/156/2020\|EPI_ISL_443248\|2020-03- | ···················· | ···················· | ····················· | ···················· | ···················· | ···················· |
| B | 2228 | hCoV-19/Singapore/157/2020\|EPI_ISL_443249\|2020-03- | ···················· | ···················· | ····················· | ···················· | ···················· | ···················· |
| B | 2229 | hCoV-19/USA/MN-UMGC-2/2020\|EPI_ISL_443253\|2020-04- | ···················· | ···················· | ····················· | ···················· | ···················· | ···················· |
| B | 2230 | hCoV-19/USA/MN-UMGC-3/2020\|EPI_ISL_443254\|2020-04- | ···················· | ···················· | ····················· | ···················· | ···················· | ···················· |
| B | 2231 | hCoV-19/USA/MN-UMGC-4/2020\|EPI_ISL_443255\|2020-03- | ···················· | ···················· | ····················· | ···················· | ···················· | ···················· |
| B | 2233 | hCoV-19/France/IDF-4791/2020\|EPI_ISL_443258\|2020-0 | ···················· | ···················· | ····················· | ···················· | ···················· | ···················· |
| B | 2234 | hCoV-19/France/IDF-4793/2020\|EPI_ISL_443259\|2020-0 | ···················· | ···················· | ····················· | ···················· | ···················· | ···················· |
| B | 2235 | hCoV-19/France/IDF-4843/2020\|EPI_ISL_443260\|2020-0 | ···················· | ···················· | ····················· | ···················· | ···················· | ···················· |
| B | 2236 | hCoV-19/France/BFC-5011/2020\|EPI_ISL_443261\|2020-0 | ···················· | ···················· | ····················· | ···················· | ···················· | ···················· |
| B | 2237 | hCoV-19/France/BFC-5012/2020\|EPI_ISL_443262\|2020-0 | ···················· | ···················· | ····················· | ···················· | ···················· | ···················· |
| B | 2238 | hCoV-19/France/BFC-5017/2020\|EPI_ISL_443263\|2020-0 | ···················· | ···················· | ····················· | ···················· | ···················· | ···················· |
| B | 2239 | hCoV-19/France/BFC-5019/2020\|EPI_ISL_443264\|2020-0 | ···················· | ···················· | ····················· | ···················· | ···················· | ···················· |
| B | 2240 | hCoV-19/France/BRE-5310/2020\|EPI_ISL_443265\|2020-0 | ···················· | ···················· | ····················· | ···················· | ···················· | ···················· |
| B | 2241 | hCoV-19/France/BRE-5322/2020\|EPI_ISL_443266\|2020-0 | ···················· | ···················· | ····················· | ···················· | ···················· | ···················· |
| B | 2242 | hCoV-19/France/BRE-5325/2020\|EPI_ISL_443267\|2020-0 | ···················· | ···················· | ····················· | ···················· | ···················· | ···················· |
| B | 2243 | hCoV-19/France/BRE-5336/2020\|EPI_ISL_443268\|2020-0 | ···················· | ···················· | ····················· | ···················· | ···················· | ···················· |
| B | 2244 | hCoV-19/France/BRE-5344/2020\|EPI_ISL_443269\|2020-0 | ···················· | ···················· | ····················· | ···················· | ···················· | ···················· |
| B | 2245 | hCoV-19/France/BRE-5353/2020\|EPI_ISL_443270\|2020-0 | ···················· | ···················· | ····················· | ···················· | ···················· | ···················· |
| B | 2246 | hCoV-19/France/BRE-5355/2020\|EPI_ISL_443271\|2020-0 | ···················· | ···················· | ····················· | ···················· | ···················· | ···················· |
| B | 2247 | hCoV-19/France/BRE-5359/2020\|EPI_ISL_443272\|2020-0 | ···················· | ···················· | ····················· | ···················· | ···················· | ···················· |
| B | 2248 | hCoV-19/France/BRE-5363/2020\|EPI_ISL_443273\|2020-0 | ···················· | ···················· | ····················· | ···················· | ···················· | ···················· |
| B | 2249 | hCoV-19/France/BRE-5374/2020\|EPI_ISL_443274\|2020-0 | ···················· | ···················· | ····················· | ···················· | ···················· | ···················· |
| B | 2250 | hCoV-19/France/BRE-5397/2020\|EPI_ISL_443275\|2020-0 | ···················· | ···················· | ····················· | ···················· | ···················· | ···················· |
| B | 2251 | hCoV-19/France/BRE-5400/2020\|EPI_ISL_443276\|2020-0 | ···················· | ···················· | ····················· | ···················· | ···················· | ···················· |
| B | 2252 | hCoV-19/France/BRE-5413/2020\|EPI_ISL_443277\|2020-0 | ···················· | ···················· | ····················· | ···················· | ···················· | ···················· |
| B | 2253 | hCoV-19/France/BRE-5415/2020\|EPI_ISL_443278\|2020-0 | ···················· | ···················· | ····················· | ···················· | ···················· | ···················· |
| B | 2254 | hCoV-19/France/BRE-5434/2020\|EPI_ISL_443279\|2020-0 | ···················· | ···················· | ····················· | ···················· | ···················· | ···················· |
| B | 2255 | hCoV-19/France/BRE-5447/2020\|EPI_ISL_443280\|2020-0 | ···················· | ···················· | ····················· | ···················· | ···················· | ···················· |
| B | 2256 | hCoV-19/France/BRE-5456/2020\|EPI_ISL_443281\|2020-0 | ···················· | ···················· | ····················· | ···················· | ···················· | ···················· |
| B | 2257 | hCoV-19/France/BRE-5458/2020\|EPI_ISL_443282\|2020-0 | ···················· | ···················· | ····················· | ···················· | ···················· | ···················· |
| B | 2258 | hCoV-19/France/BRE-5465/2020\|EPI_ISL_443283\|2020-0 | ···················· | ···················· | ····················· | ···················· | ···················· | ···················· |
| B | 2259 | hCoV-19/France/HDF-5650/2020\|EPI_ISL_443284\|2020-0 | ···················· | ···················· | ····················· | ···················· | ···················· | ···················· |
| B | 2260 | hCoV-19/France/HDF-5655/2020\|EPI_ISL_443285\|2020-0 | ···················· | ···················· | ····················· | ···················· | ···················· | ···················· |
| B | 2261 | hCoV-19/France/HDF-5657/2020\|EPI_ISL_443286\|2020-0 | ···················· | ····A··············· | ····················· | ···················· | ···················· | ···················· |
| B | 2262 | hCoV-19/France/HDF-5662/2020\|EPI_ISL_443287\|2020-0 | ···················· | ···················· | ····················· | ···················· | ···················· | ···················· |
| B | 2263 | hCoV-19/France/HDF-5666/2020\|EPI_ISL_443288\|2020-0 | ···················· | ···················· | ····················· | ···················· | ···················· | ···················· |
| B | 2264 | hCoV-19/France/BRE-5678/2020\|EPI_ISL_443289\|2020-0 | ···················· | ···················· | ····················· | ···················· | ···················· | ···················· |
| B | 2265 | hCoV-19/France/BRE-5681/2020\|EPI_ISL_443290\|2020-0 | ···················· | ···················· | ····················· | ···················· | ···················· | ···················· |
| B | 2266 | hCoV-19/France/BRE-5685/2020\|EPI_ISL_443291\|2020-0 | ···················· | ···················· | ····················· | ···················· | ···················· | ···················· |

| | | | | | | | | |
|---|---|---|---|---|---|---|---|---|
| B | 2267 | hCoV-19/France/BRE-5687/2020\|EPI_ISL_443292\|2020-0 | ···················· | ···················· | ···················· | ···················· | ···················· | ···················· |
| B | 2268 | hCoV-19/France/BRE-5688/2020\|EPI_ISL_443293\|2020-0 | ···················· | ···················· | ···················· | ···················· | ···················· | ···················· |
| B | 2269 | hCoV-19/France/BRE-5689/2020\|EPI_ISL_443294\|2020-0 | ···················· | ···················· | ···················· | ···················· | ···················· | ···················· |
| B | 2270 | hCoV-19/France/IDF-5847/2020\|EPI_ISL_443295\|2020-0 | ···················· | ···················· | ···················· | ···················· | ···················· | ···················· |
| B | 2271 | hCoV-19/France/IDF-5848/2020\|EPI_ISL_443296\|2020-0 | ···················· | ···················· | ···················· | ···················· | ···················· | ···················· |
| B | 2272 | hCoV-19/France/IDF-5851/2020\|EPI_ISL_443298\|2020-0 | ···················· | ···················· | ···················· | ···················· | ···················· | ···················· |
| B | 2273 | hCoV-19/France/IDF-5853/2020\|EPI_ISL_443299\|2020-0 | ···················· | ···················· | ···················· | ···················· | ···················· | ···················· |
| B | 2274 | hCoV-19/England/BRIS-1249DD/2020\|EPI_ISL_443891\|20 | ···················· | ···················· | ···················· | ···················· | ···················· | ···················· |
| B | 2275 | hCoV-19/England/BRIS-124A43/2020\|EPI_ISL_443895\|20 | ···················· | ···················· | ···················· | ···················· | ···················· | ···················· |
| B | 2276 | hCoV-19/USA/IL-NM088/2020\|EPI_ISL_444578\|2020-03-1 | ···················· | ···················· | ···················· | ···················· | ···················· | ···················· |
| B | 2277 | hCoV-19/USA/IL-NM089/2020\|EPI_ISL_444579\|2020-03-1 | ···················· | ···················· | ···················· | ···················· | ···················· | ···················· |
| B | 2278 | hCoV-19/USA/IL-NM090/2020\|EPI_ISL_444580\|2020-03-1 | ···················· | ···················· | ···················· | ···················· | ···················· | ···················· |
| B | 2280 | hCoV-19/USA/IL-NM095/2020\|EPI_ISL_444583\|2020-03-1 | ···················· | ···················· | ···················· | ···················· | ···················· | ···················· |
| B | 2281 | hCoV-19/USA/IL-NM097/2020\|EPI_ISL_444585\|2020-03-1 | ···················· | ···················· | ···················· | ···················· | ···················· | ···················· |
| B | 2282 | hCoV-19/USA/IL-NM098/2020\|EPI_ISL_444586\|2020-03-2 | ···················· | ···················· | ···················· | ···················· | ···················· | ···················· |
| B | 2283 | hCoV-19/USA/IL-NM0101/2020\|EPI_ISL_444587\|2020-03- | ···················· | ···················· | ···················· | ···················· | ···················· | ···················· |
| B | 2284 | hCoV-19/USA/IL-NM0103/2020\|EPI_ISL_444588\|2020-03- | ···················· | ···················· | ···················· | ···················· | ···················· | ···················· |
| B | 2285 | hCoV-19/USA/IL-NM0104/2020\|EPI_ISL_444589\|2020-03- | ···················· | ···················· | ···················· | ···················· | ···················· | ···················· |
| B | 2286 | hCoV-19/USA/IL-NM0105/2020\|EPI_ISL_444590\|2020-03- | ···················· | ···················· | ···················· | ···················· | ···················· | ···················· |
| B | 2287 | hCoV-19/USA/IL-NM0106/2020\|EPI_ISL_444591\|2020-03- | ···················· | ···················· | ···················· | ···················· | ···················· | ···················· |
| B | 2288 | hCoV-19/USA/IL-NM0107/2020\|EPI_ISL_444592\|2020-03- | ···················· | ···················· | ···················· | ···················· | ···················· | ···················· |
| B | 2289 | hCoV-19/USA/IL-NM0108/2020\|EPI_ISL_444593\|2020-03- | ···················· | ···················· | ···················· | ···················· | ···················· | ···················· |
| B | 2290 | hCoV-19/USA/IL-NM0109/2020\|EPI_ISL_444594\|2020-03- | ···················· | ···················· | ···················· | ···················· | ···················· | ···················· |
| B | 2291 | hCoV-19/USA/IL-NM0111/2020\|EPI_ISL_444595\|2020-03- | ···················· | ···················· | ···················· | ···················· | ···················· | ···················· |
| B | 2292 | hCoV-19/USA/IL-NM0112/2020\|EPI_ISL_444596\|2020-03- | ···················· | ···················· | ···················· | ···················· | ···················· | ···················· |
| B | 2293 | hCoV-19/USA/IL-NM0113/2020\|EPI_ISL_444597\|2020-03- | ···················· | ···················· | ···················· | ···················· | ···················· | ···················· |
| B | 2294 | hCoV-19/USA/IL-NM0114/2020\|EPI_ISL_444598\|2020-03- | ···················· | ···················· | ···················· | ···················· | ···················· | ···················· |
| B | 2295 | hCoV-19/USA/IL-NM0116/2020\|EPI_ISL_444599\|2020-03- | ···················· | ···················· | ···················· | ···················· | ···················· | ···················· |
| B | 2296 | hCoV-19/Wales/PHWC-2C7BE/2020\|EPI_ISL_445910\|2020- | ···················· | ···················· | ···················· | ···················· | ···················· | ···················· |
| B | 2297 | hCoV-19/Wales/PHWC-2C806/2020\|EPI_ISL_445915\|2020- | ···················· | ···················· | ···················· | ···················· | ···················· | ···················· |
| B | 2298 | hCoV-19/Wales/PHWC-2C815/2020\|EPI_ISL_445916\|2020- | ···················· | ···················· | ···················· | ···················· | ···················· | ···················· |
| B | 2299 | hCoV-19/Hong Kong/HKU-905a/2020\|EPI_ISL_450408\|202 | ···················· | ···················· | ···················· | ···················· | ···················· | ···················· |
| B | 2300 | hCoV-19/Hong Kong/HKU-911a/2020\|EPI_ISL_450409\|202 | ···················· | ···················· | ···················· | ···················· | ···················· | ···················· |
| B | 2301 | hCoV-19/Hong Kong/HKU-913a/2020\|EPI_ISL_450410\|202 | ···················· | ·A·················· | ···················· | ···················· | ···················· | ···················· |
| B | 2302 | hCoV-19/Hong Kong/HKU-915a/2020\|EPI_ISL_450411\|202 | ···················· | ·A·················· | ···················· | ···················· | ···················· | ···················· |
| B | 2303 | hCoV-19/Hong Kong/HKU-904a/2020\|EPI_ISL_450412\|202 | ···················· | ···················· | ···················· | ···················· | ···················· | ···················· |
| B | 2304 | hCoV-19/Henan/HN03/2020\|EPI_ISL_450442\|2020-02-24 | ···················· | ···················· | ···················· | ···················· | ···················· | ···················· |
| B | 2317 | hCoV-19/Australia/VIC1024/2020\|EPI_ISL_430531\|2020 | ···················· | ···················· | ···················· | ···················· | ···················· | ·····S·············· |
| B | 2319 | hCoV-19/Australia/VIC1037/2020\|EPI_ISL_430533\|2020 | ···················· | ···················· | ···················· | ···················· | ···················· | ···················· |
| B | 2321 | hCoV-19/Australia/VIC1041/2020\|EPI_ISL_430536\|2020 | ···················· | ···················· | ···················· | ···················· | ···················· | ···················· |
| B | 2322 | hCoV-19/Australia/VIC1042/2020\|EPI_ISL_430538\|2020 | ···················· | ···················· | ···················· | ···················· | ···················· | ···················· |
| B | 2323 | hCoV-19/Australia/VIC1043/2020\|EPI_ISL_430539\|2020 | ···················· | ···················· | ···················· | ···················· | ···················· | ···················· |
| B | 2324 | hCoV-19/Australia/VIC978/2020\|EPI_ISL_430542\|2020- | ···················· | ···················· | ···················· | ···················· | ···················· | ···················· |
| B | 2325 | hCoV-19/Australia/VIC982/2020\|EPI_ISL_430543\|2020- | ···················· | ···················· | ···················· | ···················· | ···················· | ···················· |
| B | 2326 | hCoV-19/Australia/VIC983/2020\|EPI_ISL_430545\|2020- | ···················· | ···················· | ···················· | ···················· | ···················· | ···················· |
| B | 2327 | hCoV-19/Australia/VIC1045/2020\|EPI_ISL_430546\|2020 | ···················· | ···················· | ···················· | ···················· | ···················· | ···················· |
| B | 2330 | hCoV-19/Australia/VIC1046/2020\|EPI_ISL_430550\|2020 | ···················· | ···················· | ···················· | ···················· | ···················· | ···················· |
| B | 2331 | hCoV-19/Australia/VIC989/2020\|EPI_ISL_430551\|2020- | ···················· | ···················· | ···················· | ···················· | ···················· | ···················· |
| B | 2334 | hCoV-19/Australia/VIC985/2020\|EPI_ISL_430554\|2020- | ···················· | ···················· | ···················· | ···················· | ···················· | ···················· |
| B | 2335 | hCoV-19/Australia/VIC990/2020\|EPI_ISL_430556\|2020- | ···················· | ···················· | ···················· | ···················· | ···················· | ···················· |
| B | 2341 | hCoV-19/Australia/VIC1112/2020\|EPI_ISL_430573\|2020 | ···················· | ···················· | ···················· | ···················· | ···················· | ···················· |
| B | 2345 | hCoV-19/Australia/VIC1115/2020\|EPI_ISL_430577\|2020 | ···················· | ···················· | ···················· | ···················· | ···················· | ···················· |
| B | 2346 | hCoV-19/Australia/VIC1062/2020\|EPI_ISL_430578\|2020 | ···················· | ···················· | ···················· | ···················· | ···················· | ···················· |
| B | 2347 | hCoV-19/Australia/VIC1080/2020\|EPI_ISL_430579\|2020 | ···················· | ···················· | ···················· | ···················· | ···················· | ···················· |
| B | 2356 | hCoV-19/England/CAMB-7E119/2020\|EPI_ISL_434041\|202 | ···················· | ···················· | ···················· | ···················· | ···················· | ···················· |
| B | 2357 | hCoV-19/England/CAMB-7E155/2020\|EPI_ISL_434045\|202 | ···················· | ···················· | ···················· | ···················· | ···················· | ···················· |
| B | 2358 | hCoV-19/England/CAMB-7E164/2020\|EPI_ISL_434046\|202 | ···················· | ···················· | ···················· | ···················· | ···················· | ···················· |
| B | 2359 | hCoV-19/England/CAMB-7E1BF/2020\|EPI_ISL_434047\|202 | ···················· | ···················· | ···················· | ···················· | ···················· | ···················· |
| B | 2360 | hCoV-19/England/CAMB-7E1DD/2020\|EPI_ISL_434049\|202 | ···················· | ···················· | ···················· | ···················· | ···················· | ···················· |
| B | 2361 | hCoV-19/England/CAMB-7E1FB/2020\|EPI_ISL_434051\|202 | ···················· | ···················· | ···················· | ···················· | ···················· | ···················· |
| B | 2362 | hCoV-19/England/CAMB-7E234/2020\|EPI_ISL_434054\|202 | ···················· | ···················· | ···················· | ···················· | ···················· | ···················· |
| B | 2363 | hCoV-19/USA/UT-0368/2020\|EPI_ISL_435356\|2020-03-29 | ···················· | ···················· | ···················· | ···················· | ···················· | ···················· |
| B | 2364 | hCoV-19/USA/UT-0522/2020\|EPI_ISL_435359\|2020-04-03 | ···················· | ···················· | ···················· | ···················· | ···················· | ···················· |
| B | 2365 | hCoV-19/USA/UT-0539/2020\|EPI_ISL_435372\|2020-04-02 | ···················· | ···················· | ···················· | ···················· | ···················· | ···················· |
| B | 2366 | hCoV-19/USA/UT-0542/2020\|EPI_ISL_435375\|2020-04-02 | ···················· | ···················· | ···················· | ···················· | ···················· | ···················· |
| B | 2367 | hCoV-19/USA/UT-0543/2020\|EPI_ISL_435376\|2020-04-02 | ···················· | ···················· | ···················· | ···················· | ···················· | ···················· |
| B | 2368 | hCoV-19/USA/UT-0544/2020\|EPI_ISL_435377\|2020-04-02 | ···················· | ···················· | ···················· | ···················· | ···················· | ···················· |
| B | 2369 | hCoV-19/USA/UT-0678/2020\|EPI_ISL_435379\|2020-04-05 | ···················· | ···················· | ···················· | ···················· | ···················· | ···················· |
| B | 2370 | hCoV-19/USA/UT-0883/2020\|EPI_ISL_435384\|2020-04-06 | ···················· | ···················· | ···················· | ···················· | ···················· | ···················· |
| B | 2371 | hCoV-19/USA/UT-0888/2020\|EPI_ISL_435389\|2020-04-06 | ···················· | ···················· | ···················· | ···················· | ···················· | ···················· |
| B | 2374 | hCoV-19/England/CAMB-7957C/2020\|EPI_ISL_441200\|202 | ···················· | ···················· | ···················· | ···················· | ···················· | ···················· |
| B | 2375 | hCoV-19/England/CAMB-795D6/2020\|EPI_ISL_441201\|202 | ···················· | ···················· | ···················· | ···················· | ···················· | ···················· |
| B | 2376 | hCoV-19/England/CAMB-78C1E/2020\|EPI_ISL_441202\|202 | ···················· | ···················· | ···················· | ···················· | ···················· | ···················· |
| B | 2377 | hCoV-19/England/CAMB-792EE/2020\|EPI_ISL_441203\|202 | ···················· | ···················· | ···················· | ···················· | ···················· | ···················· |
| B | 2378 | hCoV-19/England/CAMB-77EDD/2020\|EPI_ISL_441204\|202 | ···················· | ···················· | ···················· | ···················· | ···················· | ···················· |
| B | 2379 | hCoV-19/England/CAMB-7A98E/2020\|EPI_ISL_441205\|202 | ···················· | ···················· | ···················· | ···················· | ···················· | ···················· |
| B | 2380 | hCoV-19/England/CAMB-77F43/2020\|EPI_ISL_441206\|202 | ···················· | ···················· | ···················· | ···················· | ···················· | ···················· |
| B | 2381 | hCoV-19/England/CAMB-790F3/2020\|EPI_ISL_441207\|202 | ···················· | ···················· | ···················· | ···················· | ···················· | ···················· |

| | | | | | | | |
|---|---|---|---|---|---|---|---|
| B | 2382 hCoV-19/England/CAMB-7B668/2020\|EPI_ISL_441208\|202 | .................... | .................... | .................... | .................... | .................... | .................... |
| B | 2383 hCoV-19/England/CAMB-7963D/2020\|EPI_ISL_441209\|202 | .................... | .................... | .................... | .................... | .................... | .................... |
| B | 2384 hCoV-19/England/CAMB-79248/2020\|EPI_ISL_441210\|202 | .................... | .................... | .................... | .................... | .................... | .................... |
| B | 2385 hCoV-19/England/CAMB-798FB/2020\|EPI_ISL_441211\|202 | .................... | .................... | .................... | .................... | .................... | .................... |
| B | 2386 hCoV-19/England/CAMB-7962E/2020\|EPI_ISL_441213\|202 | .................... | .................... | .................... | .................... | .................... | .................... |
| B | 2387 hCoV-19/England/CAMB-796B5/2020\|EPI_ISL_441214\|202 | .................... | .................... | .................... | .................... | .................... | .................... |
| B | 2388 hCoV-19/England/CAMB-796F1/2020\|EPI_ISL_441215\|202 | .................... | .................... | .................... | .................... | .................... | .................... |
| B | 2389 hCoV-19/England/CAMB-78D39/2020\|EPI_ISL_441216\|202 | .................... | .................... | .................... | .................... | .................... | .................... |
| B | 2390 hCoV-19/England/CAMB-797C1/2020\|EPI_ISL_441217\|202 | .................... | .................... | .................... | .................... | .................... | .................... |
| B | 2391 hCoV-19/England/CAMB-7B61D/2020\|EPI_ISL_441218\|202 | .................... | .................... | .................... | .................... | .................... | .................... |
| B | 2392 hCoV-19/England/CAMB-78CA5/2020\|EPI_ISL_441220\|202 | .................... | .................... | .................... | .................... | .................... | .................... |
| B | 2393 hCoV-19/England/CAMB-7A872/2020\|EPI_ISL_441221\|202 | .................... | .................... | .................... | .................... | .................... | .................... |
| B | 2394 hCoV-19/England/CAMB-7915A/2020\|EPI_ISL_441222\|202 | .................... | .................... | .................... | .................... | .................... | .................... |
| B | 2395 hCoV-19/England/CAMB-7A283/2020\|EPI_ISL_441223\|202 | .................... | .................... | .................... | .................... | .................... | .................... |
| B | 2396 hCoV-19/England/CAMB-79DFC/2020\|EPI_ISL_441224\|202 | .................... | .................... | .................... | .................... | .................... | .................... |
| B | 2397 hCoV-19/England/CAMB-7904E/2020\|EPI_ISL_441225\|202 | .................... | .................... | .................... | .................... | .................... | .................... |
| B | 2398 hCoV-19/England/CAMB-79B98/2020\|EPI_ISL_441226\|202 | .................... | .................... | .................... | .................... | .................... | .................... |
| B | 2399 hCoV-19/England/CAMB-79C3B/2020\|EPI_ISL_441227\|202 | .................... | .................... | .................... | .................... | .................... | .................... |
| B | 2400 hCoV-19/England/CAMB-7A247/2020\|EPI_ISL_441228\|202 | .................... | .................... | .................... | .................... | .................... | .................... |
| B | 2401 hCoV-19/England/CAMB-77FE9/2020\|EPI_ISL_441229\|202 | .................... | .................... | .................... | .................... | .................... | .................... |
| B | 2402 hCoV-19/England/CAMB-7A229/2020\|EPI_ISL_441231\|202 | .................... | .................... | .................... | .................... | .................... | .................... |
| B | 2403 hCoV-19/England/CAMB-78E54/2020\|EPI_ISL_441232\|202 | .................... | .................... | .................... | .................... | .................... | .................... |
| B | 2404 hCoV-19/England/CAMB-7A9AC/2020\|EPI_ISL_441234\|202 | .................... | .................... | .................... | .................... | .................... | .................... |
| B | 2405 hCoV-19/England/CAMB-7A168/2020\|EPI_ISL_441236\|202 | .................... | .................... | .................... | .................... | .................... | .................... |
| B | 2406 hCoV-19/England/CAMB-78DFD/2020\|EPI_ISL_441237\|202 | .................... | .................... | .................... | .................... | .................... | .................... |
| B | 2407 hCoV-19/England/CAMB-7AA21/2020\|EPI_ISL_441239\|202 | .................... | .................... | .................... | .................... | .................... | .................... |
| B | 2408 hCoV-19/England/CAMB-78D84/2020\|EPI_ISL_441240\|202 | .................... | .................... | .................... | .................... | .................... | .................... |
| B | 2409 hCoV-19/England/CAMB-7A7B1/2020\|EPI_ISL_441242\|202 | .................... | .................... | .................... | .................... | .................... | .................... |
| B | 2410 hCoV-19/England/CAMB-77F52/2020\|EPI_ISL_441243\|202 | .................... | .................... | .................... | .................... | .................... | .................... |
| B | 2411 hCoV-19/England/CAMB-7A12C/2020\|EPI_ISL_441244\|202 | .................... | .................... | .................... | .................... | .................... | .................... |
| B | 2412 hCoV-19/England/CAMB-79F6F/2020\|EPI_ISL_441268\|202 | .................... | .................... | .................... | .................... | .................... | .................... |
| B | 2413 hCoV-19/England/CAMB-7A809/2020\|EPI_ISL_441269\|202 | .................... | .................... | .................... | .................... | .................... | .................... |
| B | 2414 hCoV-19/England/CAMB-79002/2020\|EPI_ISL_441270\|202 | .................... | .................... | .................... | .................... | .................... | .................... |
| B | 2415 hCoV-19/England/CAMB-7A793/2020\|EPI_ISL_441271\|202 | .................... | .................... | .................... | .................... | .................... | .................... |
| B | 2416 hCoV-19/England/CAMB-78EEB/2020\|EPI_ISL_441272\|202 | .................... | .................... | .................... | .................... | .................... | .................... |
| B | 2417 hCoV-19/England/CAMB-79FAB/2020\|EPI_ISL_441274\|202 | .................... | .................... | .................... | .................... | .................... | .................... |
| B | 2418 hCoV-19/England/CAMB-7A1E0/2020\|EPI_ISL_441278\|202 | .................... | .................... | .................... | .................... | .................... | .................... |
| B | 2419 hCoV-19/England/CAMB-78E45/2020\|EPI_ISL_441280\|202 | .................... | .................... | .................... | .................... | .................... | .................... |
| B | 2420 hCoV-19/England/CAMB-78E27/2020\|EPI_ISL_441281\|202 | .................... | .................... | .................... | .................... | .................... | .................... |
| B | 2421 hCoV-19/England/CAMB-79E17/2020\|EPI_ISL_441282\|202 | .................... | .................... | .................... | .................... | .................... | .................... |
| B | 2422 hCoV-19/England/CAMB-7A177/2020\|EPI_ISL_441283\|202 | .................... | .................... | .................... | .................... | .................... | .................... |
| B | 2423 hCoV-19/England/CAMB-79C0E/2020\|EPI_ISL_441284\|202 | .................... | .................... | .................... | .................... | .................... | .................... |
| B | 2424 hCoV-19/England/CAMB-7B589/2020\|EPI_ISL_441286\|202 | .................... | .................... | .................... | .................... | .................... | .................... |
| B | 2425 hCoV-19/England/CAMB-78E36/2020\|EPI_ISL_441288\|202 | .................... | .................... | .................... | .................... | .................... | .................... |
| B | 2426 hCoV-19/England/CAMB-7A14A/2020\|EPI_ISL_441289\|202 | .................... | .................... | .................... | .................... | .................... | .................... |
| B | 2427 hCoV-19/England/CAMB-7A0E3/2020\|EPI_ISL_441290\|202 | .................... | .................... | .................... | .................... | .................... | .................... |
| B | 2428 hCoV-19/England/CAMB-79DB0/2020\|EPI_ISL_441292\|202 | .................... | .................... | .................... | .................... | .................... | .................... |
| B | 2429 hCoV-19/England/CAMB-7A353/2020\|EPI_ISL_441293\|202 | .................... | .................... | .................... | .................... | .................... | .................... |
| B | 2430 hCoV-19/England/CAMB-7A7DF/2020\|EPI_ISL_441294\|202 | .................... | .................... | .................... | .................... | .................... | .................... |
| B | 2431 hCoV-19/England/CAMB-79C2C/2020\|EPI_ISL_441295\|202 | .................... | .................... | .................... | .................... | .................... | .................... |
| B | 2432 hCoV-19/England/CAMB-79C77/2020\|EPI_ISL_441296\|202 | .................... | .................... | .................... | .................... | .................... | .................... |
| B | 2433 hCoV-19/England/CAMB-79E08/2020\|EPI_ISL_441297\|202 | .................... | .................... | .................... | .................... | .................... | .................... |
| B | 2434 hCoV-19/England/CAMB-79D29/2020\|EPI_ISL_441298\|202 | .................... | .................... | .................... | .................... | .................... | .................... |
| B | 2435 hCoV-19/England/CAMB-79DED/2020\|EPI_ISL_441299\|202 | .................... | .................... | .................... | .................... | .................... | .................... |
| B | 2436 hCoV-19/England/SHEF-D1F74/2020\|EPI_ISL_442502\|202 | .................... | .................... | .................... | .................... | .................... | .................... |
| B | 2437 hCoV-19/England/SHEF-D21D7/2020\|EPI_ISL_442506\|202 | .................... | .................... | .................... | .................... | .................... | .................... |
| B | 2438 hCoV-19/England/SHEF-D2957/2020\|EPI_ISL_442510\|202 | .................... | .................... | .................... | .................... | .................... | .................... |
| B | 2439 hCoV-19/Iran/KHGRC-1.1-IPI-8206/2020\|EPI_ISL_44252 | .................... | .................... | .................... | .................... | .................... | .................... |
| B | 2440 hCoV-19/England/CAMB-818D2/2020\|EPI_ISL_442524\|202 | .................... | .................... | .................... | .................... | .................... | .................... |
| B | 2441 hCoV-19/England/CAMB-81B60/2020\|EPI_ISL_442526\|202 | .................... | .................... | .................... | .................... | .................... | .................... |
| B | 2442 hCoV-19/England/CAMB-814B0/2020\|EPI_ISL_442527\|202 | .................... | .................... | .................... | .................... | .................... | .................... |
| B | 2443 hCoV-19/England/CAMB-8183C/2020\|EPI_ISL_442528\|202 | .................... | .................... | .................... | .................... | .................... | .................... |
| B | 2444 hCoV-19/England/CAMB-818E1/2020\|EPI_ISL_442529\|202 | .................... | .................... | .................... | .................... | .................... | .................... |
| B | 2445 hCoV-19/England/CAMB-81702/2020\|EPI_ISL_442531\|202 | .................... | .................... | .................... | .................... | .................... | .................... |
| B | 2446 hCoV-19/England/CAMB-819EE/2020\|EPI_ISL_442532\|202 | .................... | .................... | .................... | .................... | .................... | .................... |
| B | 2447 hCoV-19/England/CAMB-7E850/2020\|EPI_ISL_442533\|202 | .................... | .................... | .................... | .................... | .................... | .................... |
| B | 2448 hCoV-19/England/CAMB-81878/2020\|EPI_ISL_442534\|202 | .................... | .................... | .................... | .................... | .................... | .................... |
| B | 2449 hCoV-19/England/CAMB-817C6/2020\|EPI_ISL_442535\|202 | .................... | .................... | .................... | .................... | .................... | .................... |
| B | 2450 hCoV-19/England/CAMB-8076D/2020\|EPI_ISL_442536\|202 | .................... | .................... | .................... | .................... | .................... | .................... |
| B | 2451 hCoV-19/England/CAMB-7E7EA/2020\|EPI_ISL_442537\|202 | .................... | .................... | .................... | .................... | .................... | .................... |
| B | 2452 hCoV-19/England/CAMB-817B7/2020\|EPI_ISL_442538\|202 | .................... | .................... | .................... | .................... | .................... | .................... |
| B | 2453 hCoV-19/England/CAMB-81E0D/2020\|EPI_ISL_442539\|202 | .................... | .................... | .................... | .................... | .................... | .................... |
| B | 2454 hCoV-19/England/CAMB-81BF7/2020\|EPI_ISL_442541\|202 | .................... | .................... | .................... | .................... | .................... | .................... |
| B | 2455 hCoV-19/England/CAMB-81465/2020\|EPI_ISL_442542\|202 | .................... | .................... | .................... | .................... | .................... | .................... |
| B | 2456 hCoV-19/England/CAMB-7E683/2020\|EPI_ISL_442543\|202 | .................... | .................... | .................... | .................... | .................... | .................... |
| B | 2457 hCoV-19/England/CAMB-7E823/2020\|EPI_ISL_442545\|202 | .................... | .................... | .................... | .................... | .................... | .................... |
| B | 2458 hCoV-19/England/CAMB-7E8E7/2020\|EPI_ISL_442546\|202 | .................... | .................... | .................... | .................... | .................... | .................... |
| B | 2459 hCoV-19/England/CAMB-8174E/2020\|EPI_ISL_442548\|202 | .................... | .................... | .................... | .................... | .................... | .................... |

| | | | | | | | | |
|---|---|---|---|---|---|---|---|---|
| B | 2460 | hCoV-19/England/CAMB-7EA4B/2020\|EPI_ISL_442553\|202 | ···················· | ···················· | ····················· | ···················· | ···················· | ···················· |
| B | 2461 | hCoV-19/England/CAMB-8074F/2020\|EPI_ISL_442556\|202 | ···················· | ···················· | ····················· | ···················· | ···················· | ···················· |
| B | 2462 | hCoV-19/England/CAMB-81517/2020\|EPI_ISL_442558\|202 | ···················· | ···················· | ····················· | ···················· | ···················· | ···················· |
| B | 2463 | hCoV-19/England/CAMB-81C9A/2020\|EPI_ISL_442559\|202 | ···················· | ···················· | ····················· | ···················· | ···················· | ···················· |
| B | 2464 | hCoV-19/England/CAMB-81B9D/2020\|EPI_ISL_442561\|202 | ···················· | ···················· | ····················· | ···················· | ···················· | ···················· |
| B | 2465 | hCoV-19/England/CAMB-806AC/2020\|EPI_ISL_442562\|202 | ···················· | ···················· | ····················· | ···················· | ···················· | ···················· |
| B | 2466 | hCoV-19/England/CAMB-816F6/2020\|EPI_ISL_442563\|202 | ···················· | ···················· | ····················· | ···················· | ···················· | ···················· |
| B | 2467 | hCoV-19/England/CAMB-81799/2020\|EPI_ISL_442564\|202 | ···················· | ···················· | ····················· | ···················· | ···················· | ···················· |
| B | 2468 | hCoV-19/England/CAMB-81CA9/2020\|EPI_ISL_442565\|202 | ···················· | ···················· | ····················· | ···················· | ···················· | ···················· |
| B | 2469 | hCoV-19/England/CAMB-81D6A/2020\|EPI_ISL_442567\|202 | ···················· | ···················· | ····················· | ···················· | ···················· | ···················· |
| B | 2470 | hCoV-19/England/CAMB-816D8/2020\|EPI_ISL_442569\|202 | ···················· | ···················· | ····················· | ···················· | ···················· | ···················· |
| B | 2471 | hCoV-19/England/CAMB-81B15/2020\|EPI_ISL_442570\|202 | ···················· | ···················· | ····················· | ···················· | ···················· | ···················· |
| B | 2472 | hCoV-19/England/CAMB-819C0/2020\|EPI_ISL_442572\|202 | ···················· | ···················· | ····················· | ···················· | ···················· | ···················· |
| B | 2473 | hCoV-19/England/CAMB-81474/2020\|EPI_ISL_442573\|202 | ···················· | ···················· | ····················· | ···················· | ···················· | ···················· |
| B | 2474 | hCoV-19/England/CAMB-7E8D8/2020\|EPI_ISL_442574\|202 | ···················· | ···················· | ····················· | ···················· | ···················· | ···················· |
| B | 2475 | hCoV-19/England/CAMB-81C21/2020\|EPI_ISL_442575\|202 | ···················· | ···················· | ····················· | ···················· | ···················· | ···················· |
| B | 2476 | hCoV-19/England/CAMB-7EA2D/2020\|EPI_ISL_442599\|202 | ···················· | ···················· | ····················· | ···················· | ···················· | ···················· |
| B | 2477 | hCoV-19/England/BRIS-1255AB/2020\|EPI_ISL_443800\|20 | ···················· | ···················· | ····················· | ···················· | ···················· | ···················· |
| B | 2478 | hCoV-19/England/BRIS-1249A0/2020\|EPI_ISL_443801\|20 | ···················· | ···················· | ····················· | ···················· | ···················· | ···················· |
| B | 2479 | hCoV-19/England/BRIS-125769/2020\|EPI_ISL_443802\|20 | ···················· | ···················· | ····················· | ···················· | ···················· | ···················· |
| B | 2480 | hCoV-19/England/BRIS-125295/2020\|EPI_ISL_443803\|20 | ···················· | ···················· | ····················· | ···················· | ···················· | ···················· |
| B | 2481 | hCoV-19/England/BRIS-1255E7/2020\|EPI_ISL_443804\|20 | ···················· | ···················· | ····················· | ···················· | ···················· | ···················· |
| B | 2482 | hCoV-19/England/BRIS-1251C5/2020\|EPI_ISL_443805\|20 | ···················· | ···················· | ····················· | ···················· | ···················· | ···················· |
| B | 2483 | hCoV-19/England/BRIS-125D67/2020\|EPI_ISL_443807\|20 | ···················· | ···················· | ····················· | ···················· | ···················· | ···················· |
| B | 2484 | hCoV-19/England/BRIS-12512F/2020\|EPI_ISL_443808\|20 | ···················· | ···················· | ····················· | ···················· | ···················· | ···················· |
| B | 2485 | hCoV-19/England/BRIS-124D1D/2020\|EPI_ISL_443809\|20 | ···················· | ···················· | ····················· | ···················· | ···················· | ···················· |
| B | 2486 | hCoV-19/England/BRIS-124E0B/2020\|EPI_ISL_443811\|20 | ···················· | ···················· | ····················· | ···················· | ···················· | ···················· |
| B | 2487 | hCoV-19/England/BRIS-125198/2020\|EPI_ISL_443812\|20 | ···················· | ···················· | ····················· | ···················· | ···················· | ···················· |
| B | 2488 | hCoV-19/England/BRIS-124FEA/2020\|EPI_ISL_443813\|20 | ···················· | ···················· | ····················· | ···················· | ···················· | ···················· |
| B | 2489 | hCoV-19/England/BRIS-124ADA/2020\|EPI_ISL_443815\|20 | ···················· | ···················· | ····················· | ···················· | ···················· | ···················· |
| B | 2490 | hCoV-19/England/BRIS-124F62/2020\|EPI_ISL_443816\|20 | ···················· | ···················· | ····················· | ···················· | ···················· | ···················· |
| B | 2491 | hCoV-19/England/BRIS-124876/2020\|EPI_ISL_443817\|20 | ···················· | ···················· | ····················· | ···················· | ···················· | ···················· |
| B | 2492 | hCoV-19/England/BRIS-125B21/2020\|EPI_ISL_443819\|20 | ···················· | ···················· | ····················· | ···················· | ···················· | ···················· |
| B | 2493 | hCoV-19/England/BRIS-12571E/2020\|EPI_ISL_443820\|20 | ···················· | ···················· | ····················· | ···················· | ···················· | ···················· |
| B | 2494 | hCoV-19/England/BRIS-125E0A/2020\|EPI_ISL_443821\|20 | ···················· | ···················· | ····················· | ···················· | ···················· | ···················· |
| B | 2495 | hCoV-19/England/BRIS-1249BF/2020\|EPI_ISL_443825\|20 | ···················· | ···················· | ····················· | ···················· | ···················· | ···················· |
| B | 2496 | hCoV-19/England/BRIS-125BA9/2020\|EPI_ISL_443826\|20 | ···················· | ···················· | ····················· | ···················· | ···················· | ···················· |
| B | 2497 | hCoV-19/England/BRIS-125EFB/2020\|EPI_ISL_443827\|20 | ···················· | ···················· | ····················· | ···················· | ···················· | ···················· |
| B | 2498 | hCoV-19/England/BRIS-12558D/2020\|EPI_ISL_443828\|20 | ···················· | ···················· | ····················· | ···················· | ···················· | ···················· |
| B | 2499 | hCoV-19/England/BRIS-125778/2020\|EPI_ISL_443830\|20 | ···················· | ···················· | ····················· | ···················· | ···················· | ···················· |
| B | 2500 | hCoV-19/England/BRIS-125ECE/2020\|EPI_ISL_443831\|20 | ···················· | ···················· | ····················· | ···················· | ···················· | ···················· |
| B | 2501 | hCoV-19/England/BRIS-124F44/2020\|EPI_ISL_443832\|20 | ···················· | ···················· | ····················· | ···················· | ···················· | ···················· |
| B | 2502 | hCoV-19/England/BRIS-1247D3/2020\|EPI_ISL_443833\|20 | ············T······· | ···················· | ····················· | ···················· | ···················· | ···················· |
| B | 2503 | hCoV-19/England/BRIS-1256D5/2020\|EPI_ISL_443834\|20 | ···················· | ···················· | ····················· | ···················· | ···················· | ···················· |
| B | 2504 | hCoV-19/England/BRIS-124C89/2020\|EPI_ISL_443835\|20 | ···················· | ···················· | ····················· | ···················· | ···················· | ···················· |
| B | 2505 | hCoV-19/England/BRIS-1248B2/2020\|EPI_ISL_443836\|20 | ···················· | ···················· | ····················· | ···················· | ···················· | ···R················ |
| B | 2506 | hCoV-19/England/BRIS-125F34/2020\|EPI_ISL_443838\|20 | ···················· | ···················· | ····················· | ···················· | ···················· | ···················· |
| B | 2507 | hCoV-19/England/BRIS-124A07/2020\|EPI_ISL_443839\|20 | ···················· | ····A··············· | ····················· | ···················· | ···················· | ···················· |
| B | 2508 | hCoV-19/England/BRIS-125D2B/2020\|EPI_ISL_443840\|20 | ···················· | ···················· | ····················· | ···················· | ···················· | ···················· |
| B | 2509 | hCoV-19/England/BRIS-125F16/2020\|EPI_ISL_443841\|20 | ···················· | ···················· | ····················· | ···················· | ···················· | ···················· |
| B | 2510 | hCoV-19/England/BRIS-125550/2020\|EPI_ISL_443842\|20 | ···················· | ···················· | ····················· | ···················· | ···················· | ···················· |
| B | 2511 | hCoV-19/England/BRIS-124B31/2020\|EPI_ISL_443843\|20 | ···················· | ···················· | ····················· | ···················· | ···················· | ···················· |
| B | 2512 | hCoV-19/England/BRIS-124BE6/2020\|EPI_ISL_443844\|20 | ···················· | ···················· | ····················· | ···················· | ···················· | ···················· |
| B | 2513 | hCoV-19/England/BRIS-124ECF/2020\|EPI_ISL_443846\|20 | ···················· | ···················· | ····················· | ···················· | ···················· | ···················· |
| B | 2514 | hCoV-19/England/BRIS-125787/2020\|EPI_ISL_443847\|20 | ···················· | ···················· | ····················· | ···················· | ···················· | ···················· |
| B | 2515 | hCoV-19/England/BRIS-125E46/2020\|EPI_ISL_443851\|20 | ···················· | ···················· | ····················· | ···················· | ···················· | ···················· |
| B | 2516 | hCoV-19/England/BRIS-12523B/2020\|EPI_ISL_443852\|20 | ···················· | ···················· | ····················· | ···················· | ···················· | ···················· |
| B | 2517 | hCoV-19/England/BRIS-125E73/2020\|EPI_ISL_443853\|20 | ···················· | ···················· | ····················· | ···················· | ···················· | ···················· |
| B | 2518 | hCoV-19/England/BRIS-124AF8/2020\|EPI_ISL_443854\|20 | ···················· | ···················· | ····················· | ···················· | ···················· | ···················· |
| B | 2519 | hCoV-19/England/BRIS-125D49/2020\|EPI_ISL_443855\|20 | ···················· | ···················· | ····················· | ···················· | ···················· | ···················· |
| B | 2520 | hCoV-19/England/BRIS-12520E/2020\|EPI_ISL_443856\|20 | ···················· | ···················· | ····················· | ···················· | ···················· | ···················· |
| B | 2521 | hCoV-19/England/BRIS-1250E6/2020\|EPI_ISL_443857\|20 | ···················· | ···················· | ····················· | ···················· | ···················· | ···················· |
| B | 2522 | hCoV-19/England/BRIS-125C2E/2020\|EPI_ISL_443858\|20 | ···················· | ···················· | ····················· | ···················· | ···················· | ···················· |
| B | 2523 | hCoV-19/England/BRIS-124F9F/2020\|EPI_ISL_443859\|20 | ···················· | ···················· | ····················· | ···················· | ···················· | ···················· |
| B | 2524 | hCoV-19/England/BRIS-125ABB/2020\|EPI_ISL_443860\|20 | ···················· | ···················· | ····················· | ···················· | ···················· | ···················· |
| B | 2525 | hCoV-19/England/BRIS-124CA7/2020\|EPI_ISL_443862\|20 | ···················· | ···················· | ····················· | ···················· | ···················· | ···················· |
| B | 2526 | hCoV-19/England/BRIS-124A16/2020\|EPI_ISL_443864\|20 | ···················· | ···················· | ····················· | ···················· | ···················· | ···················· |
| B | 2527 | hCoV-19/England/BRIS-125963/2020\|EPI_ISL_443865\|20 | ···················· | ···················· | ····················· | ···················· | ···················· | ···················· |
| B | 2528 | hCoV-19/England/BRIS-1258C0/2020\|EPI_ISL_443866\|20 | ···················· | ···················· | ····················· | ···················· | ···················· | ···················· |
| B | 2529 | hCoV-19/England/BRIS-1251F2/2020\|EPI_ISL_443867\|20 | ···················· | ···················· | ····················· | ···················· | ···················· | ···················· |
| B | 2530 | hCoV-19/England/BRIS-125277/2020\|EPI_ISL_443869\|20 | ···················· | ···················· | ····················· | ···················· | ···················· | ···················· |
| B | 2531 | hCoV-19/England/BRIS-12515C/2020\|EPI_ISL_443870\|20 | ···················· | ···················· | ····················· | ···················· | ···················· | ···················· |
| B | 2532 | hCoV-19/England/BRIS-124EFC/2020\|EPI_ISL_443871\|20 | ···················· | ···················· | ····················· | ···················· | ···················· | ···················· |
| B | 2533 | hCoV-19/England/BRIS-124ACB/2020\|EPI_ISL_443872\|20 | ···················· | ···················· | ····················· | ···················· | ···················· | ···················· |
| B | 2534 | hCoV-19/England/BRIS-1250D7/2020\|EPI_ISL_443873\|20 | ···················· | ···················· | ····················· | ···················· | ···················· | ···················· |
| B | 2535 | hCoV-19/England/BRIS-1258FD/2020\|EPI_ISL_443876\|20 | ···················· | ···················· | ····················· | ···················· | ···················· | ···················· |
| B | 2536 | hCoV-19/England/BRIS-1256C6/2020\|EPI_ISL_443877\|20 | ···················· | ···················· | ····················· | ···················· | ···················· | ···················· |
| B | 2537 | hCoV-19/England/BRIS-125392/2020\|EPI_ISL_443879\|20 | ···················· | ···················· | ····················· | ···················· | ···················· | ···················· |

| | | | | | | | | | |
|---|---|---|---|---|---|---|---|---|---|
| B | 2538 | hCoV-19/England/BRIS-125F61/2020\|EPI_ISL_443880\|20 | .................... | .................... | .................... | .................... | .................... | .................... | |
| B | 2539 | hCoV-19/England/BRIS-12559C/2020\|EPI_ISL_443881\|20 | .................... | .................... | .................... | .................... | .................... | .................... | |
| B | 2540 | hCoV-19/England/BRIS-124A70/2020\|EPI_ISL_443882\|20 | .................... | .................... | .................... | .................... | .................... | .................... | |
| B | 2541 | hCoV-19/England/BRIS-124E92/2020\|EPI_ISL_443883\|20 | .................... | .................... | .................... | .................... | .................... | .................... | |
| B | 2542 | hCoV-19/England/BRIS-12521D/2020\|EPI_ISL_443884\|20 | .................... | .................... | .................... | .................... | .................... | .................... | |
| B | 2543 | hCoV-19/England/BRIS-1251E3/2020\|EPI_ISL_443885\|20 | .................... | .................... | .................... | .................... | .................... | .................... | |
| B | 2544 | hCoV-19/England/BRIS-1258B1/2020\|EPI_ISL_443888\|20 | .................... | .................... | .................... | .................... | .................... | .................... | |
| B | 2545 | hCoV-19/England/BRIS-124849/2020\|EPI_ISL_443892\|20 | .................... | .................... | .................... | .................... | .................... | .................... | |
| B | 2546 | hCoV-19/England/BRIS-125CB5/2020\|EPI_ISL_443893\|20 | .................... | .................... | .................... | .................... | .................... | .................... | |
| B | 2547 | hCoV-19/England/BRIS-125F07/2020\|EPI_ISL_443894\|20 | .................... | .................... | .................... | .................... | .................... | .................... | |
| B | 2548 | hCoV-19/England/BRIS-1255BA/2020\|EPI_ISL_443896\|20 | .................... | .................... | .................... | .................... | .................... | .................... | |
| B | 2549 | hCoV-19/England/BRIS-124D0E/2020\|EPI_ISL_443897\|20 | .................... | .................... | .................... | .................... | .................... | .................... | |
| B | 2550 | hCoV-19/England/BRIS-125D0D/2020\|EPI_ISL_443898\|20 | .................... | .................... | .................... | .................... | .................... | .................... | |
| B | 2551 | hCoV-19/England/BRIS-125A7F/2020\|EPI_ISL_443899\|20 | .................... | .................... | .................... | .................... | .................... | .................... | |
| B | 2552 | hCoV-19/Wales/PHWC-2DDF7/2020\|EPI_ISL_446025\|2020- | .................... | .................... | .................... | .................... | .................... | .................... | |
| B | 2553 | hCoV-19/Wales/PHWC-2DE21/2020\|EPI_ISL_446027\|2020- | .................... | .................... | .................... | .................... | .................... | .................... | |
| B | 2554 | hCoV-19/Wales/PHWC-2DE6D/2020\|EPI_ISL_446031\|2020- | .................... | .................... | .................... | .................... | .................... | .................... | |
| B | 2555 | hCoV-19/Wales/PHWC-2DEF4/2020\|EPI_ISL_446038\|2020- | .................... | .................... | .................... | .................... | .................... | .................... | |
| B | 2556 | hCoV-19/Wales/PHWC-2DF00/2020\|EPI_ISL_446039\|2020- | .................... | .................... | .................... | .................... | .................... | .................... | |
| B | 2557 | hCoV-19/Wales/PHWC-2E29D/2020\|EPI_ISL_446080\|2020- | .................... | .................... | .................... | .................... | .................... | .................... | |
| B | 2558 | hCoV-19/Wales/PHWC-2E2CA/2020\|EPI_ISL_446083\|2020- | .................... | .................... | .................... | .................... | .................... | .................... | |
| B | 2559 | hCoV-19/Wales/PHWC-2E2F7/2020\|EPI_ISL_446086\|2020- | .................... | .................... | .................... | .................... | .................... | .................... | |
| B | 2560 | hCoV-19/Wales/PHWC-2E9F2/2020\|EPI_ISL_446098\|2020- | .................... | .................... | .................... | .................... | .................... | .................... | |
| B | 2561 | hCoV-19/England/CAMB-7C166/2020\|EPI_ISL_434001\|202 | .................... | .................... | .................... | .................... | .................... | .................... | |
| B | 2562 | hCoV-19/England/CAMB-7C175/2020\|EPI_ISL_434002\|202 | .................... | .................... | .................... | .................... | .................... | .................... | |
| B | 2563 | hCoV-19/England/CAMB-7C1C0/2020\|EPI_ISL_434007\|202 | .................... | .................... | .................... | .................... | .................... | .................... | |
| B | 2564 | hCoV-19/England/CAMB-7C1EE/2020\|EPI_ISL_434009\|202 | .................... | .................... | .................... | .................... | .................... | .................... | |
| B | 2565 | hCoV-19/England/CAMB-7C1FD/2020\|EPI_ISL_434010\|202 | .................... | .................... | .................... | .................... | .................... | .................... | |
| B | 2566 | hCoV-19/England/CAMB-7C218/2020\|EPI_ISL_434012\|202 | .................... | .................... | .................... | .................... | .................... | .................... | |
| B | 2567 | hCoV-19/England/CAMB-7C227/2020\|EPI_ISL_434013\|202 | .................... | .................... | .................... | .................... | .................... | .................... | |
| B | 2568 | hCoV-19/England/CAMB-7C236/2020\|EPI_ISL_434014\|202 | .................... | .................... | .................... | .................... | .................... | .................... | |
| B | 2569 | hCoV-19/England/CAMB-7C245/2020\|EPI_ISL_434015\|202 | .................... | .................... | .................... | .................... | .................... | .................... | |
| B | 2570 | hCoV-19/England/CAMB-7C290/2020\|EPI_ISL_434020\|202 | .................... | ...................A | .................... | .................... | .................... | .................... | 1 |
| B | 2571 | hCoV-19/England/CAMB-7C2AF/2020\|EPI_ISL_434021\|202 | .................... | .................... | .................... | .................... | .................... | .................... | |
| B | 2572 | hCoV-19/England/CAMB-7C2BE/2020\|EPI_ISL_434022\|202 | .................... | .................... | .................... | .................... | .................... | .................... | |
| B | 2573 | hCoV-19/England/CAMB-7C2DC/2020\|EPI_ISL_434024\|202 | .................... | .................... | .................... | .................... | .................... | .................... | |
| B | 2574 | hCoV-19/England/CAMB-7C315/2020\|EPI_ISL_434027\|202 | .................... | .................... | .................... | .................... | .................... | .................... | |
| B | 2575 | hCoV-19/England/CAMB-7C333/2020\|EPI_ISL_434029\|202 | .................... | .................... | .................... | .................... | .................... | .................... | |
| B | 2576 | hCoV-19/England/CAMB-7C360/2020\|EPI_ISL_434032\|202 | .................... | .................... | .................... | .................... | .................... | .................... | |
| B | 2577 | hCoV-19/England/CAMB-7C38E/2020\|EPI_ISL_434034\|202 | .................... | .................... | .................... | .................... | .................... | .................... | |
| B | 2578 | hCoV-19/England/CAMB-7C39D/2020\|EPI_ISL_434035\|202 | .................... | .................... | .................... | .................... | .................... | .................... | |
| B | 2579 | hCoV-19/England/CAMB-7C3AC/2020\|EPI_ISL_434036\|202 | .................... | .................... | .................... | .................... | .................... | .................... | |
| B | 2580 | hCoV-19/England/CAMB-7C3CA/2020\|EPI_ISL_434038\|202 | .................... | .................... | .................... | .................... | .................... | .................... | |
| B | 2581 | hCoV-19/England/CAMB-7C3E8/2020\|EPI_ISL_434039\|202 | .................... | ...G................ | .................... | .................... | .................... | .................... | |
| B | 2582 | hCoV-19/USA/WA-S303/2020\|EPI_ISL_434063\|2020-04-03 | .................... | .................... | .................... | .................... | .................... | .................... | |
| B | 2583 | hCoV-19/USA/WA-S304/2020\|EPI_ISL_434064\|2020-04-05 | .................... | .................... | .................... | .................... | .................... | .................... | |
| B | 2584 | hCoV-19/USA/WA-S305/2020\|EPI_ISL_434065\|2020-04-01 | .................... | .................... | .................... | .................... | .................... | .................... | |
| B | 2585 | hCoV-19/USA/WA-S306/2020\|EPI_ISL_434066\|2020-04-02 | .................... | .................... | .................... | .................... | .................... | .................... | |
| B | 2586 | hCoV-19/USA/WA-S308/2020\|EPI_ISL_434068\|2020-04-02 | .................... | .................... | .................... | .................... | .................... | .................... | |
| B | 2587 | hCoV-19/USA/WA-S309/2020\|EPI_ISL_434069\|2020-04-03 | .................... | .................... | .................... | .................... | .................... | .................... | |
| B | 2588 | hCoV-19/USA/WA-S310/2020\|EPI_ISL_434070\|2020-04-01 | .................... | .................... | .................... | .................... | .................... | .................... | |
| B | 2589 | hCoV-19/USA/WA-S311/2020\|EPI_ISL_434071\|2020-04-04 | .................... | .................... | .................... | .................... | .................... | .................... | |
| B | 2590 | hCoV-19/USA/WA-S312/2020\|EPI_ISL_434072\|2020-04-01 | .................... | .................... | .................... | .................... | .................... | .................... | |
| B | 2591 | hCoV-19/USA/WA-S313/2020\|EPI_ISL_434073\|2020-04-02 | .................... | .................... | .................... | .................... | .................... | .................... | |
| B | 2592 | hCoV-19/USA/WA-S314/2020\|EPI_ISL_434074\|2020-04-03 | .................... | .................... | .................... | .................... | .................... | .................... | |
| B | 2593 | hCoV-19/USA/WA-S315/2020\|EPI_ISL_434075\|2020-04-02 | .................... | .................... | .................... | .................... | .................... | .................... | |
| B | 2594 | hCoV-19/USA/WA-S316/2020\|EPI_ISL_434076\|2020-04-01 | .................... | .................... | .................... | .................... | .................... | .................... | |
| B | 2595 | hCoV-19/USA/WA-S317/2020\|EPI_ISL_434077\|2020-04-04 | .................... | .................... | .................... | .................... | .................... | .................... | |
| B | 2596 | hCoV-19/USA/WA-S319/2020\|EPI_ISL_434079\|2020-04-04 | .................... | .................... | .................... | .................... | .................... | .................... | |
| B | 2597 | hCoV-19/USA/WA-S320/2020\|EPI_ISL_434080\|2020-04-01 | .................... | .................... | .................... | .................... | .................... | .................... | |
| B | 2598 | hCoV-19/USA/WA-S321/2020\|EPI_ISL_434081\|2020-03-30 | .................... | .................... | .................... | .................... | .................... | .................... | |
| B | 2599 | hCoV-19/USA/WA-S322/2020\|EPI_ISL_434082\|2020-03-31 | .................... | .................... | .................... | .................... | .................... | .................... | |
| B | 2600 | hCoV-19/USA/WA-S323/2020\|EPI_ISL_434083\|2020-04-02 | .................... | .................... | .................... | .................... | .................... | .................... | |
| B | 2601 | hCoV-19/USA/WA-S324/2020\|EPI_ISL_434084\|2020-04-01 | .................... | .................... | .................... | .................... | .................... | .................... | |
| B | 2602 | hCoV-19/USA/WA-S325/2020\|EPI_ISL_434085\|2020-04-03 | .................... | .................... | .................... | .................... | .................... | .................... | |
| B | 2603 | hCoV-19/USA/WA-S326/2020\|EPI_ISL_434086\|2020-04-01 | .................... | .................... | .................... | .................... | .................... | .................... | |
| B | 2604 | hCoV-19/USA/WA-S327/2020\|EPI_ISL_434087\|2020-04-05 | .................... | .................... | .................... | .................... | .................... | .................... | |
| B | 2605 | hCoV-19/USA/WA-S328/2020\|EPI_ISL_434088\|2020-04-03 | .................... | .................... | .................... | .................... | .................... | .................... | |
| B | 2606 | hCoV-19/USA/WA-S329/2020\|EPI_ISL_434089\|2020-04-01 | .................... | .................... | .................... | .................... | .................... | .................... | |
| B | 2607 | hCoV-19/USA/WA-S330/2020\|EPI_ISL_434090\|2020-04-02 | .................... | .................... | .................... | .................... | .................... | .................... | |
| B | 2608 | hCoV-19/USA/WA-S331/2020\|EPI_ISL_434091\|2020-04-02 | .................... | .................... | .................... | .................... | .................... | .................... | |
| B | 2609 | hCoV-19/USA/WA-S332/2020\|EPI_ISL_434092\|2020-04-02 | .................... | .................... | .................... | .................... | .................... | .................... | |
| B | 2610 | hCoV-19/USA/WA-S333/2020\|EPI_ISL_434093\|2020-04-04 | .................... | .................... | .................... | .................... | .................... | .................... | |
| B | 2611 | hCoV-19/USA/WA-S334/2020\|EPI_ISL_434094\|2020-04-04 | .................... | .................... | .................... | .................... | .................... | .................... | |
| B | 2612 | hCoV-19/USA/WA-S335/2020\|EPI_ISL_434095\|2020-04-02 | .................... | .................... | .................... | .................... | .................... | .................... | |
| B | 2613 | hCoV-19/USA/WA-S336/2020\|EPI_ISL_434096\|2020-04-03 | .................... | .................... | .................... | .................... | .................... | .................... | |
| B | 2614 | hCoV-19/USA/WA-S337/2020\|EPI_ISL_434097\|2020-04-01 | .................... | .................... | .................... | .................... | .................... | .................... | |
| B | 2615 | hCoV-19/USA/WA-S338/2020\|EPI_ISL_434098\|2020-04-02 | .................... | .................... | .................... | .................... | .................... | .................... | |

| | | | | | | | | |
|---|---|---|---|---|---|---|---|---|
| B | 2616 | hCoV-19/USA/WA-S339/2020\|EPI_ISL_434099\|2020-04-02 | ···················· | ···················· | ····················· | ···················· | ···················· | ···················· |
| B | 2617 | hCoV-19/Vietnam/VNHN_022/2020\|EPI_ISL_435303\|2020- | ···················· | ···················· | ····················· | ···················· | ···················· | ···················· |
| B | 2618 | hCoV-19/Vietnam/VNHN_026/2020\|EPI_ISL_435305\|2020- | ···················· | ···················· | ····················· | ···················· | ···················· | ···················· |
| B | 2619 | hCoV-19/Vietnam/VNHN_0299/2020\|EPI_ISL_435308\|2020 | ···················· | ···················· | ····················· | ···················· | ···················· | ···················· |
| B | 2620 | hCoV-19/Vietnam/VNHN_0300/2020\|EPI_ISL_435310\|2020 | ···················· | ···················· | ····················· | ···················· | ···················· | ···················· |
| B | 2621 | hCoV-19/Vietnam/VNHN_0418/2020\|EPI_ISL_435311\|2020 | ···················· | ···················· | ····················· | ···················· | ···················· | ···················· |
| B | 2622 | hCoV-19/Vietnam/VNHN_0419/2020\|EPI_ISL_435312\|2020 | ···················· | ···················· | ····················· | ···················· | ···················· | ···················· |
| B | 2623 | hCoV-19/Vietnam/VNHN_0762/2020\|EPI_ISL_435313\|2020 | ···················· | ···················· | ····················· | ···················· | ···················· | ···················· |
| B | 2624 | hCoV-19/Vietnam/VNHN_0764/2020\|EPI_ISL_435314\|2020 | ···················· | ···················· | ····················· | ···················· | ···················· | ···················· |
| B | 2625 | hCoV-19/Vietnam/VNHN_0837/2020\|EPI_ISL_435315\|2020 | ···················· | ···················· | ····················· | ···················· | ···················· | ···················· |
| B | 2626 | hCoV-19/Vietnam/VNHN_0985/2020\|EPI_ISL_435316\|2020 | ···················· | ···················· | ····················· | ···················· | ···················· | ···················· |
| B | 2627 | hCoV-19/Vietnam/VNHN_01097/2020\|EPI_ISL_435317\|202 | ···················· | ·············A······ | ····················· | ···················· | ···················· | ···················· |
| B | 2628 | hCoV-19/USA/UT-0106/2020\|EPI_ISL_435350\|2020-03-12 | ···················· | ···················· | ····················· | ···················· | ···················· | ···················· |
| B | 2629 | hCoV-19/USA/UT-0353/2020\|EPI_ISL_435353\|2020-03-27 | ···················· | ···················· | ····················· | ···················· | ···················· | ···················· |
| B | 2630 | hCoV-19/USA/UT-0521/2020\|EPI_ISL_435358\|2020-04-03 | ···················· | ···················· | ····················· | ···················· | ···················· | ···················· |
| B | 2631 | hCoV-19/USA/UT-0524/2020\|EPI_ISL_435360\|2020-04-03 | ···················· | ···················· | ····················· | ···················· | ···················· | ···················· |
| B | 2632 | hCoV-19/USA/UT-0526/2020\|EPI_ISL_435362\|2020-04-03 | ···················· | ···················· | ····················· | ···················· | ···················· | ···················· |
| B | 2633 | hCoV-19/USA/UT-0527/2020\|EPI_ISL_435363\|2020-04-03 | ···················· | ···················· | ····················· | ···················· | ···················· | ···················· |
| B | 2634 | hCoV-19/USA/UT-0534/2020\|EPI_ISL_435368\|2020-04-02 | ···················· | ···················· | ····················· | ···················· | ···················· | ···················· |
| B | 2635 | hCoV-19/USA/UT-0535/2020\|EPI_ISL_435369\|2020-04-03 | ···················· | ···················· | ····················· | ···················· | ···················· | ···················· |
| B | 2636 | hCoV-19/USA/UT-0891/2020\|EPI_ISL_435390\|2020-04-06 | ···················· | ···················· | ····················· | ···················· | ···················· | ···················· |
| B | 2637 | hCoV-19/USA/UT-0894/2020\|EPI_ISL_435391\|2020-04-05 | ···················· | ···················· | ····················· | ···················· | ···················· | ···················· |
| B | 2638 | hCoV-19/USA/UT-0896/2020\|EPI_ISL_435393\|2020-04-07 | ···················· | ···················· | ····················· | ···················· | ···················· | ···················· |
| B | 2639 | hCoV-19/USA/WI-GMF-01455/2020\|EPI_ISL_435394\|2020- | ···················· | ···················· | ····················· | ···················· | ···················· | ···················· |
| B | 2640 | hCoV-19/USA/WI-GMF-01481/2020\|EPI_ISL_435395\|2020- | ···················· | ···················· | ····················· | ···················· | ···················· | ···················· |
| B | 2641 | hCoV-19/USA/WI-GMF-01535/2020\|EPI_ISL_435397\|2020- | ···················· | ···················· | ····················· | ···················· | ···················· | ···················· |
| B | 2642 | hCoV-19/USA/WI-GMF-01551/2020\|EPI_ISL_435398\|2020- | ···················· | ···················· | ····················· | ···················· | ···················· | ···················· |
| B | 2643 | hCoV-19/USA/WI-GMF-01591/2020\|EPI_ISL_435399\|2020- | ···················· | ···················· | ····················· | ···················· | ···················· | ···················· |
| B | 2644 | hCoV-19/USA/WI-UW-306/2020\|EPI_ISL_436600\|2020-04- | ···················· | ···················· | ····················· | ···················· | ···················· | ···················· |
| B | 2645 | hCoV-19/USA/WI-UW-307/2020\|EPI_ISL_436601\|2020-04- | ···················· | ···················· | ····················· | ···················· | ···················· | ···················· |
| B | 2646 | hCoV-19/USA/WI-UW-308/2020\|EPI_ISL_436602\|2020-04- | ···················· | ···················· | ····················· | ···················· | ···················· | ···················· |
| B | 2647 | hCoV-19/USA/WI-UW-309/2020\|EPI_ISL_436603\|2020-04- | ···················· | ···················· | ····················· | ···················· | ···················· | ···················· |
| B | 2648 | hCoV-19/USA/WI-UW-310/2020\|EPI_ISL_436604\|2020-04- | ···················· | ···················· | ····················· | ···················· | ···················· | ···················· |
| B | 2649 | hCoV-19/USA/WI-UW-311/2020\|EPI_ISL_436605\|2020-04- | ···················· | ···················· | ····················· | ···················· | ···················· | ···················· |
| B | 2650 | hCoV-19/USA/WI-UW-312/2020\|EPI_ISL_436606\|2020-04- | ···················· | ···················· | ····················· | ···················· | ···················· | ···················· |
| B | 2651 | hCoV-19/USA/WI-UW-313/2020\|EPI_ISL_436607\|2020-04- | ···················· | ···················· | ····················· | ···················· | ···················· | ···················· |
| B | 2652 | hCoV-19/USA/WI-UW-314/2020\|EPI_ISL_436608\|2020-04- | ···················· | ···················· | ····················· | ···················· | ···················· | ···················· |
| B | 2653 | hCoV-19/USA/WI-UW-315/2020\|EPI_ISL_436609\|2020-04- | ···················· | ···················· | ····················· | ···················· | ···················· | ···················· |
| B | 2654 | hCoV-19/USA/WI-UW-316/2020\|EPI_ISL_436610\|2020-04- | ···················· | ···················· | ····················· | ···················· | ···················· | ···················· |
| B | 2655 | hCoV-19/USA/WI-UW-318/2020\|EPI_ISL_436612\|2020-04- | ···················· | ···················· | ····················· | ···················· | ···················· | ···················· |
| B | 2656 | hCoV-19/USA/WI-UW-319/2020\|EPI_ISL_436613\|2020-04- | ···················· | ···················· | ····················· | ···················· | ···················· | ···················· |
| B | 2657 | hCoV-19/USA/WI-UW-320/2020\|EPI_ISL_436614\|2020-04- | ···················· | ···················· | ····················· | ···················· | ···················· | ···················· |
| B | 2658 | hCoV-19/USA/WI-UW-321/2020\|EPI_ISL_436615\|2020-04- | ···················· | ···················· | ····················· | ···················· | ···················· | ···················· |
| B | 2659 | hCoV-19/USA/WI-UW-322/2020\|EPI_ISL_436616\|2020-04- | ···················· | ···················· | ····················· | ···················· | ···················· | ···················· |
| B | 2660 | hCoV-19/USA/WI-UW-323/2020\|EPI_ISL_436617\|2020-04- | ···················· | ···················· | ····················· | ···················· | ···················· | ···················· |
| B | 2661 | hCoV-19/USA/WI-UW-324/2020\|EPI_ISL_436618\|2020-04- | ···················· | ···················· | ····················· | ···················· | ···················· | ···················· |
| B | 2662 | hCoV-19/USA/WI-UW-325/2020\|EPI_ISL_436619\|2020-04- | ···················· | ·A·················· | ····················· | ···················· | ···················· | ···················· |
| B | 2663 | hCoV-19/USA/WI-UW-326/2020\|EPI_ISL_436620\|2020-04- | ···················· | ···················· | ····················· | ···················· | ···················· | ···················· |
| B | 2664 | hCoV-19/USA/WI-UW-327/2020\|EPI_ISL_436621\|2020-04- | ···················· | ···················· | ····················· | ···················· | ···················· | ···················· |
| B | 2665 | hCoV-19/USA/WI-UW-328/2020\|EPI_ISL_436622\|2020-04- | ···················· | ···················· | ····················· | ···················· | ···················· | ···················· |
| B | 2666 | hCoV-19/USA/WI-UW-329/2020\|EPI_ISL_436623\|2020-04- | ···················· | ···················· | ····················· | ···················· | ···················· | ···················· |
| B | 2667 | hCoV-19/USA/WI-UW-331/2020\|EPI_ISL_436625\|2020-04- | ···················· | ···················· | ····················· | ···················· | ···················· | ···················· |
| B | 2668 | hCoV-19/USA/WI-UW-332/2020\|EPI_ISL_436626\|2020-04- | ···················· | ···················· | ····················· | ···················· | ···················· | ···················· |
| B | 2669 | hCoV-19/USA/WI-UW-333/2020\|EPI_ISL_436627\|2020-03- | ···················· | ···················· | ····················· | ···················· | ···················· | ···················· |
| B | 2670 | hCoV-19/USA/WI-UW-334/2020\|EPI_ISL_436628\|2020-03- | ···················· | ···················· | ····················· | ···················· | ···················· | ···················· |
| B | 2671 | hCoV-19/USA/WI-UW-335/2020\|EPI_ISL_436629\|2020-03- | ···················· | ···················· | ····················· | ···················· | ···················· | ···················· |
| B | 2672 | hCoV-19/USA/WI-UW-336/2020\|EPI_ISL_436630\|2020-03- | ···················· | ···················· | ····················· | ···················· | ···················· | ···················· |
| B | 2673 | hCoV-19/USA/WI-UW-337/2020\|EPI_ISL_436631\|2020-03- | ···················· | ···················· | ····················· | ···················· | ···················· | ···················· |
| B | 2674 | hCoV-19/USA/WI-UW-338/2020\|EPI_ISL_436632\|2020-03- | ···················· | ···················· | ····················· | ···················· | ···················· | ···················· |
| B | 2675 | hCoV-19/USA/WI-UW-340/2020\|EPI_ISL_436634\|2020-04- | ···················· | ···················· | ····················· | ···················· | ···················· | ···················· |
| B | 2676 | hCoV-19/USA/WI-UW-341/2020\|EPI_ISL_436635\|2020-04- | ···················· | ···················· | ····················· | ···················· | ···················· | ···················· |
| B | 2677 | hCoV-19/USA/WI-UW-342/2020\|EPI_ISL_436636\|2020-04- | ···················· | ···················· | ····················· | ···················· | ···················· | ···················· |
| B | 2678 | hCoV-19/USA/WI-UW-343/2020\|EPI_ISL_436637\|2020-04- | ···················· | ···················· | ····················· | ···················· | ···················· | ···················· |
| B | 2679 | hCoV-19/USA/WI-UW-344/2020\|EPI_ISL_436638\|2020-04- | ···················· | ···················· | ····················· | ···················· | ···················· | ···················· |
| B | 2680 | hCoV-19/USA/WI-UW-346/2020\|EPI_ISL_436640\|2020-04- | ···················· | ···················· | ····················· | ···················· | ···················· | ···················· |
| B | 2681 | hCoV-19/USA/CA-CZB-1000/2020\|EPI_ISL_436641\|2020-0 | ···················· | ···················· | ····················· | ···················· | ···················· | ···················· |
| B | 2682 | hCoV-19/USA/CA-CZB-1001/2020\|EPI_ISL_436642\|2020-0 | ···················· | ···················· | ····················· | ···················· | ···················· | ···················· |
| B | 2683 | hCoV-19/USA/CA-CZB-1002/2020\|EPI_ISL_436643\|2020-0 | ···················· | ···················· | ····················· | ···················· | ···················· | ···················· |
| B | 2684 | hCoV-19/USA/CA-CZB-1003/2020\|EPI_ISL_436644\|2020-0 | ···················· | ···················· | ····················· | ···················· | ···················· | ···················· |
| B | 2685 | hCoV-19/USA/CA-CZB-1004/2020\|EPI_ISL_436645\|2020-0 | ···················· | ···················· | ····················· | ···················· | ···················· | ···················· |
| B | 2686 | hCoV-19/USA/CA-CZB-1005/2020\|EPI_ISL_436646\|2020-0 | ···················· | ···················· | ····················· | ···················· | ···················· | ···················· |
| B | 2687 | hCoV-19/USA/CA-CZB-1006/2020\|EPI_ISL_436647\|2020-0 | ···················· | ···················· | ····················· | ···················· | ···················· | ···················· |
| B | 2688 | hCoV-19/USA/CA-CZB-1007/2020\|EPI_ISL_436648\|2020-0 | ···················· | ···················· | ····················· | ···················· | ···················· | ···················· |
| B | 2689 | hCoV-19/USA/CA-CZB-1011/2020\|EPI_ISL_436652\|2020-0 | ···················· | ···················· | ····················· | ···················· | ···················· | ···················· |
| B | 2690 | hCoV-19/USA/CA-CZB-1012/2020\|EPI_ISL_436653\|2020-0 | ···················· | ···················· | ····················· | ···················· | ···················· | ···················· |
| B | 2691 | hCoV-19/USA/CA-CZB-1013/2020\|EPI_ISL_436654\|2020-0 | ···················· | ···················· | ····················· | ···················· | ···················· | ···················· |
| B | 2692 | hCoV-19/USA/CA-CZB-1014/2020\|EPI_ISL_436655\|2020-0 | ···················· | ···················· | ····················· | ···················· | ···················· | ···················· |
| B | 2693 | hCoV-19/USA/CA-CZB-1015/2020\|EPI_ISL_436656\|2020-0 | ···················· | ···················· | ····················· | ···················· | ···················· | ···················· |

```
B 2694 hCoV-19/USA/CA-CZB-1016/2020|EPI_ISL_436657|2020-0 ···················· ···················· ····················· ···················· ···················· ····················
B 2695 hCoV-19/USA/CA-CZB-1017/2020|EPI_ISL_436658|2020-0 ···················· ···················· ····················· ···················· ···················· ····················
B 2696 hCoV-19/USA/CA-CZB-1018/2020|EPI_ISL_436659|2020-0 ···················· ···················· ····················· ···················· ···················· ····················
B 2697 hCoV-19/USA/CA-CZB-1019/2020|EPI_ISL_436660|2020-0 ···················· ···················· ····················· ···················· ···················· ····················
B 2698 hCoV-19/USA/CA-CZB-1020/2020|EPI_ISL_436661|2020-0 ···················· ···················· ····················· ···················· ···················· ····················
B 2699 hCoV-19/USA/CA-CZB-1021/2020|EPI_ISL_436662|2020-0 ···················· ···················· ····················· ···················· ···················· ····················
B 2700 hCoV-19/USA/CA-CZB-1022/2020|EPI_ISL_436663|2020-0 ···················· ···················· ····················· ···················· ···················· ····················
B 2701 hCoV-19/USA/CA-CZB-1023/2020|EPI_ISL_436664|2020-0 ···················· ···················· ····················· ···················· ···················· ····················
B 2702 hCoV-19/USA/CA-CZB-1024/2020|EPI_ISL_436665|2020-0 ···················· ···················· ····················· ···················· ···················· ····················
B 2703 hCoV-19/USA/CA-CZB-1025/2020|EPI_ISL_436666|2020-0 ···················· ···················· ····················· ···················· ···················· ····················
B 2704 hCoV-19/USA/CA-CZB-1026/2020|EPI_ISL_436667|2020-0 ···················· ···················· ····················· ···················· ···················· ····················
B 2705 hCoV-19/USA/CA-CZB-1028/2020|EPI_ISL_436669|2020-0 ···················· ···················· ····················· ···················· ···················· ····················
B 2706 hCoV-19/USA/CA-CZB-1029/2020|EPI_ISL_436670|2020-0 ···················· ···················· ····················· ···················· ···················· ····················
B 2707 hCoV-19/USA/CA-CZB-1030/2020|EPI_ISL_436671|2020-0 ···················· ···················· ····················· ···················· ···················· ····················
B 2708 hCoV-19/USA/CA-CZB-1031/2020|EPI_ISL_436672|2020-0 ···················· ···················· ····················· ···················· ···················· ····················
B 2709 hCoV-19/USA/CA-CZB-1032/2020|EPI_ISL_436673|2020-0 ···················· ···················· ····················· ···················· ···················· ····················
B 2710 hCoV-19/USA/CA-CZB-1033/2020|EPI_ISL_436674|2020-0 ···················· ···················· ····················· ···················· ···················· ····················
B 2711 hCoV-19/USA/CA-CZB-1034/2020|EPI_ISL_436675|2020-0 ···················· ···················· ····················· ···················· ···················· ····················
B 2712 hCoV-19/USA/CA-CZB-1035/2020|EPI_ISL_436676|2020-0 ···················· ···················· ····················· ···················· ···················· ····················
B 2713 hCoV-19/USA/CA-CZB-1036/2020|EPI_ISL_436677|2020-0 ···················· ···················· ····················· ···················· ···················· ····················
B 2714 hCoV-19/USA/CA-CZB-1037/2020|EPI_ISL_436678|2020-0 ···················· ···················· ····················· ···················· ···················· ····················
B 2715 hCoV-19/USA/CA-CZB-1038/2020|EPI_ISL_436679|2020-0 ···················· ···················· ····················· ···················· ···················· ····················
B 2716 hCoV-19/USA/CA-CZB-1039/2020|EPI_ISL_436680|2020-0 ···················· ···················· ····················· ···················· ···················· ····················
B 2717 hCoV-19/USA/CA-CZB-1040/2020|EPI_ISL_436681|2020-0 ···················· ···················· ····················· ···················· ···················· ····················
B 2718 hCoV-19/USA/CA-CZB-1041/2020|EPI_ISL_436682|2020-0 ···················· ···················· ····················· ···················· ···················· ····················
B 2719 hCoV-19/USA/CA-CZB-1042/2020|EPI_ISL_436683|2020-0 ···················· ···················· ····················· ···················· ···················· ····················
B 2720 hCoV-19/South Africa/KRISP-04/2020|EPI_ISL_436684| ···················· ···················· ····················· ···················· ···················· ····················
B 2721 hCoV-19/South Africa/KRISP-045/2020|EPI_ISL_436686 ···················· ···················· ····················· ···················· ···················· ····················
B 2722 hCoV-19/Austria/CeMM0034/2020|EPI_ISL_437923|2020- ···················· ···················· ····················· ···················· ···················· ····················
B 2723 hCoV-19/Austria/CeMM0035/2020|EPI_ISL_437924|2020- ···················· ···················· ····················· ···················· ···················· ····················
B 2724 hCoV-19/Austria/CeMM0036/2020|EPI_ISL_437925|2020- ···················· ···················· ····················· ···················· ···················· ····················
B 2725 hCoV-19/Austria/CeMM0037/2020|EPI_ISL_437926|2020- ···················· ···················· ····················· ···················· ···················· ····················
B 2726 hCoV-19/Austria/CeMM0038/2020|EPI_ISL_437927|2020- ···················· ···················· ····················· ···················· ···················· ····················
B 2727 hCoV-19/Austria/CeMM0039/2020|EPI_ISL_437928|2020- ···················· ···················· ····················· ···················· ···················· ····················
B 2728 hCoV-19/Austria/CeMM0041/2020|EPI_ISL_437929|2020- ···················· ···················· ····················· ···················· ···················· ····················
B 2729 hCoV-19/Austria/CeMM0042/2020|EPI_ISL_437930|2020- ···················· ···················· ····················· ···················· ···················· ····················
B 2730 hCoV-19/Austria/CeMM0043/2020|EPI_ISL_437931|2020- ···················· ···················· ····················· ···················· ···················· ····················
B 2731 hCoV-19/Austria/CeMM0045/2020|EPI_ISL_437932|2020- ···················· ···················· ····················· ···················· ···················· ····················
B 2732 hCoV-19/Austria/CeMM0047/2020|EPI_ISL_437933|2020- ···················· ···················· ····················· ···················· ···················· ····················
B 2733 hCoV-19/Austria/CeMM0048/2020|EPI_ISL_437934|2020- ···················· ···················· ····················· ···················· ···················· ····················
B 2734 hCoV-19/Austria/CeMM0052/2020|EPI_ISL_437935|2020- ···················· ···················· ····················· ···················· ···················· ····················
B 2735 hCoV-19/Austria/CeMM0053/2020|EPI_ISL_437936|2020- ···················· ···················· ····················· ···················· ···················· ····················
B 2736 hCoV-19/Austria/CeMM0055/2020|EPI_ISL_437937|2020- ···················· ···················· ····················· ···················· ···················· ····················
B 2737 hCoV-19/Austria/CeMM0058/2020|EPI_ISL_437938|2020- ···················· ···················· ····················· ···················· ···················· ····················
B 2738 hCoV-19/Austria/CeMM0067/2020|EPI_ISL_437939|2020- ···················· ···················· ····················· ···················· ···················· ····················
B 2739 hCoV-19/Austria/CeMM0068/2020|EPI_ISL_437940|2020- ···················· ···················· ····················· ···················· ···················· ····················
B 2740 hCoV-19/Austria/CeMM0071/2020|EPI_ISL_437941|2020- ···················· ···················· ····················· ···················· ···················· ···················N 1
B 2741 hCoV-19/Austria/CeMM0073/2020|EPI_ISL_437942|2020- ···················· ···················· ····················· ···················· ···················· ····················
B 2742 hCoV-19/Austria/CeMM0074/2020|EPI_ISL_437943|2020- ···················· ···················· ····················· ···················· ···················· ····················
B 2743 hCoV-19/Austria/CeMM0076/2020|EPI_ISL_437944|2020- ···················· ···A················ ····················· ···················· ···················· ····················
B 2744 hCoV-19/Austria/CeMM0077/2020|EPI_ISL_437945|2020- ···················· ···················· ····················· ···················· ···················· ····················
B 2745 hCoV-19/Austria/CeMM0078/2020|EPI_ISL_437946|2020- ···················· ···················· ····················· ···················· ···················· ····················
B 2746 hCoV-19/Austria/CeMM0080/2020|EPI_ISL_437947|2020- ···················· ···················· ····················· ···················· ···················· ····················
B 2747 hCoV-19/Austria/CeMM0083/2020|EPI_ISL_437948|2020- ···················· ···················· ····················· ···················· ···················· ····················
B 2748 hCoV-19/Austria/CeMM0084/2020|EPI_ISL_437949|2020- ···················· ···················· ····················· ···················· ···················· ····················
B 2749 hCoV-19/Austria/CeMM0086/2020|EPI_ISL_437950|2020- ···················· ···················· ····················· ···················· ···················· ····················
B 2750 hCoV-19/Austria/CeMM0088/2020|EPI_ISL_437951|2020- ···················· ···················· ····················· ···················· ···················· ····················
B 2751 hCoV-19/Austria/CeMM0089/2020|EPI_ISL_437952|2020- ···················· ···················· ····················· ···················· ···················· ····················
B 2752 hCoV-19/Austria/CeMM0090/2020|EPI_ISL_437953|2020- ···················· ···················· ····················· ···················· ···················· ···············N····
B 2753 hCoV-19/Austria/CeMM0091/2020|EPI_ISL_437954|2020- ···················· ···················· ····················· ···················· ···················· ····················
B 2754 hCoV-19/Austria/CeMM0094/2020|EPI_ISL_437955|2020- ···················· ···················· ····················· ···················· ···················· ····················
B 2755 hCoV-19/Austria/CeMM0095/2020|EPI_ISL_437956|2020- ···················· ···················· ····················· ···················· ···················· ····················
B 2756 hCoV-19/Austria/CeMM0096/2020|EPI_ISL_437957|2020- ···················· ···················· ····················· ···················· ···················· ·······N············
B 2757 hCoV-19/Austria/CeMM0097/2020|EPI_ISL_437958|2020- ···················· ···················· ····················· ···················· ···················· ····················
B 2758 hCoV-19/Austria/CeMM0098/2020|EPI_ISL_437959|2020- ···················· ···················· ····················· ···················· ···················· ····················
B 2759 hCoV-19/Austria/CeMM0099/2020|EPI_ISL_437960|2020- ···················· ···················· ····················· ···················· ···················· ····················
B 2760 hCoV-19/Austria/CeMM0100/2020|EPI_ISL_437961|2020- ···················· ···················· ····················· ···················· ···················· ····················
B 2761 hCoV-19/Austria/CeMM0101/2020|EPI_ISL_437962|2020- ···················· ···················· ····················· ···················· ···················· ····················
B 2762 hCoV-19/Austria/CeMM0102/2020|EPI_ISL_437963|2020- ···················· ···················· ····················· ···················· ···················· ····················
B 2763 hCoV-19/Austria/CeMM0103/2020|EPI_ISL_437964|2020- ···················· ···················· ····················· ···················· ···················· ····················
B 2764 hCoV-19/Austria/CeMM0104/2020|EPI_ISL_437965|2020- ···················· ···················· ····················· ···················· ···················· ····················
B 2765 hCoV-19/Austria/CeMM0106/2020|EPI_ISL_437966|2020- ···················· ···················· ····················· ···················· ···················· ····················
B 2766 hCoV-19/Austria/CeMM0108/2020|EPI_ISL_437967|2020- ···················· ·············M······ ····················· ···················· ···················· ····················
B 2767 hCoV-19/Austria/CeMM0109/2020|EPI_ISL_437968|2020- ···················· ···················· ····················· ···················· ···················· ····················
B 2768 hCoV-19/Austria/CeMM0111/2020|EPI_ISL_437969|2020- ···················· ···················· ····················· ···················· ···················· ····················
B 2769 hCoV-19/Austria/CeMM0112/2020|EPI_ISL_437970|2020- ···················· ···················· ····················· ···················· ···················· ····················
B 2770 hCoV-19/Austria/CeMM0113/2020|EPI_ISL_437971|2020- ···················· ···················· ····················· ···················· ···················· ····················
B 2771 hCoV-19/Austria/CeMM0114/2020|EPI_ISL_437972|2020- ···················· ···················· ····················· ···················· ···················· ····················
```

| | | | | | | | | | |
|---|---|---|---|---|---|---|---|---|---|
| B | 2772 | hCoV-19/Austria/CeMM0115/2020\|EPI_ISL_437973\|2020- | ···················· | ····A··············· | ···················· | ···················· | ···················· | ···················· | |
| B | 2773 | hCoV-19/Austria/CeMM0116/2020\|EPI_ISL_437974\|2020- | ···················· | ···················· | ···················· | ···················· | ···················· | ···················· | |
| B | 2774 | hCoV-19/Austria/CeMM0117/2020\|EPI_ISL_437975\|2020- | ···················· | ···················· | ···················· | ···················· | ···················· | ···················· | |
| B | 2775 | hCoV-19/Austria/CeMM0119/2020\|EPI_ISL_437976\|2020- | ···················· | ···················· | ···················· | ···················· | ···················· | ···················· | |
| B | 2776 | hCoV-19/Austria/CeMM0120/2020\|EPI_ISL_437977\|2020- | ···················· | ···················· | ···················· | ···················· | ···················· | ···················· | |
| B | 2777 | hCoV-19/Austria/CeMM0121/2020\|EPI_ISL_437978\|2020- | ···················· | ···················· | ···················· | ···················· | ···················· | ···················· | |
| B | 2778 | hCoV-19/Austria/CeMM0123/2020\|EPI_ISL_437979\|2020- | ···················· | ···················· | ···················· | ···················· | ···················· | ···················· | |
| B | 2779 | hCoV-19/Austria/CeMM0127/2020\|EPI_ISL_437983\|2020- | ···················· | ···················· | ···················· | ···················· | ···················· | ···················· | |
| B | 2780 | hCoV-19/Austria/CeMM0128/2020\|EPI_ISL_437984\|2020- | ···················· | ···················· | ···················· | ···················· | ···················· | ···················· | |
| B | 2781 | hCoV-19/Austria/CeMM0132/2020\|EPI_ISL_437985\|2020- | ···················· | ···················· | ···················· | ···················· | ···················· | ···················· | |
| B | 2782 | hCoV-19/Austria/CeMM0133/2020\|EPI_ISL_437986\|2020- | ···················· | ···················· | ···················· | ···················· | ···················· | ···················· | |
| B | 2783 | hCoV-19/Austria/CeMM0134/2020\|EPI_ISL_437987\|2020- | ···················· | ···················· | ···················· | ···················· | ···················· | ···················· | |
| B | 2784 | hCoV-19/Austria/CeMM0135/2020\|EPI_ISL_437988\|2020- | ···················· | ···················· | ···················· | ···················· | ···················· | ···················· | |
| B | 2785 | hCoV-19/Austria/CeMM0136/2020\|EPI_ISL_437989\|2020- | ···················· | ···················· | ···················· | ···················· | ···················· | ···················· | |
| B | 2786 | hCoV-19/Austria/CeMM0137/2020\|EPI_ISL_437990\|2020- | ···················· | ···················· | ···················· | ···················· | ···················· | ···················· | |
| B | 2787 | hCoV-19/Austria/CeMM0139/2020\|EPI_ISL_437991\|2020- | ···················· | ···················· | ···················· | ···················· | ···················· | ···················· | |
| B | 2788 | hCoV-19/Austria/CeMM0140/2020\|EPI_ISL_437992\|2020- | ···················· | ···················· | ···················· | ···················· | ···················· | ···················· | |
| B | 2789 | hCoV-19/Austria/CeMM0143/2020\|EPI_ISL_437993\|2020- | ···················· | ···················· | ···················· | ···················· | ···················· | ···················· | |
| B | 2790 | hCoV-19/Austria/CeMM0146/2020\|EPI_ISL_437994\|2020- | ···················· | ···················· | ···················· | ···················· | ···················· | ···················· | |
| B | 2791 | hCoV-19/Austria/CeMM0147/2020\|EPI_ISL_437995\|2020- | ···················· | ···················· | ···················· | ···················· | ···················· | ···················· | |
| B | 2792 | hCoV-19/Austria/CeMM0148/2020\|EPI_ISL_437996\|2020- | ···················· | ···················· | ···················· | ···················· | ···················· | ···················· | |
| B | 2793 | hCoV-19/Austria/CeMM0149/2020\|EPI_ISL_437997\|2020- | ···················· | ···················· | ···················· | ···················· | ···················· | ···················· | |
| B | 2794 | hCoV-19/Austria/CeMM0150/2020\|EPI_ISL_437998\|2020- | ···················· | ···················· | ···················· | ···················· | ···················· | ···················· | |
| B | 2795 | hCoV-19/Austria/CeMM0153/2020\|EPI_ISL_437999\|2020- | ···················· | ···················· | ···················· | ···················· | ···················· | ···················· | |
| B | 2796 | hCoV-19/England/CAMB-78F60/2020\|EPI_ISL_441191\|202 | ···················· | ···················· | ···················· | ···················· | ···················· | ···················· | |
| B | 2797 | hCoV-19/England/CAMB-7920C/2020\|EPI_ISL_441192\|202 | ···················· | ···················· | ···················· | ···················· | ···················· | ···················· | |
| B | 2798 | hCoV-19/England/CAMB-7966A/2020\|EPI_ISL_441194\|202 | ···················· | ···················· | ···················· | ···················· | ···················· | ···················· | |
| B | 2799 | hCoV-19/England/CAMB-7B62C/2020\|EPI_ISL_441195\|202 | ···················· | ···················· | ···················· | ···················· | ···················· | ···················· | |
| B | 2800 | hCoV-19/England/CAMB-79424/2020\|EPI_ISL_441196\|202 | ···················· | ···················· | ···················· | ···················· | ···················· | ···················· | |
| B | 2801 | hCoV-19/England/CAMB-7964C/2020\|EPI_ISL_441198\|202 | ···················· | ···················· | ···················· | ···················· | ···················· | ···················· | |
| B | 2802 | hCoV-19/Wales/PHWC-2D6ED/2020\|EPI_ISL_446014\|2020- | ···················· | ···················· | ···················· | ···················· | ···················· | ···················· | |
| B | 2803 | hCoV-19/Wales/PHWC-2DF1F/2020\|EPI_ISL_446040\|2020- | ···················· | ···················· | ···················· | ···················· | ···················· | ···················· | |
| B | 2804 | hCoV-19/Wales/PHWC-2DF6A/2020\|EPI_ISL_446042\|2020- | ···················· | ···················· | ···················· | ···················· | ···················· | ···················· | |
| B | 2805 | hCoV-19/Wales/PHWC-2DF97/2020\|EPI_ISL_446043\|2020- | ···················· | ····A··············· | ···················· | ···················· | ···················· | ···················· | |
| B | 2806 | hCoV-19/Wales/PHWC-2DFA6/2020\|EPI_ISL_446044\|2020- | ···················· | ···················· | ···················· | ···················· | ···················· | ···················· | |
| B | 2807 | hCoV-19/Wales/PHWC-2E01B/2020\|EPI_ISL_446047\|2020- | ···················· | ···················· | ···················· | ···················· | ···················· | ···················· | |
| B | 2808 | hCoV-19/Wales/PHWC-2E048/2020\|EPI_ISL_446050\|2020- | ···················· | ···················· | ···················· | ···················· | ···················· | ···················· | |
| B | 2809 | hCoV-19/Wales/PHWC-2E127/2020\|EPI_ISL_446060\|2020- | ···················· | ···················· | ···················· | ···················· | ···················· | ···················· | |
| B | 2810 | hCoV-19/Wales/PHWC-2E136/2020\|EPI_ISL_446061\|2020- | ···················· | ····A··············· | ···················· | ···················· | ···················· | ···················· | |
| B | 2811 | hCoV-19/Wales/PHWC-2E172/2020\|EPI_ISL_446064\|2020- | ···················· | ···················· | ···················· | ···················· | ···················· | ···················· | |
| B | 2812 | hCoV-19/Wales/PHWC-2E1AF/2020\|EPI_ISL_446067\|2020- | ···················· | ···················· | ···················· | ···················· | ···················· | ···················· | |
| B | 2813 | hCoV-19/Wales/PHWC-2E1BE/2020\|EPI_ISL_446068\|2020- | ···················· | ···················· | ···················· | ···················· | ···················· | ···················· | |
| B | 2814 | hCoV-19/Wales/PHWC-2E1CD/2020\|EPI_ISL_446069\|2020- | ···················· | ···················· | ···················· | ···················· | ···················· | ···················· | |
| B | 2815 | hCoV-19/Wales/PHWC-2E206/2020\|EPI_ISL_446072\|2020- | ···················· | ···················· | ···················· | ···················· | ···················· | ···················· | |
| B | 2816 | hCoV-19/Wales/PHWC-2E28E/2020\|EPI_ISL_446079\|2020- | ···················· | ···················· | ···················· | ···················· | ···················· | ···················· | |
| B | 2828 | hCoV-19/Australia/VIC942/2020\|EPI_ISL_430639\|2020- | ···················· | ···················· | ···················· | ···················· | ···················· | ···················· | |
| B | 2832 | hCoV-19/Australia/VIC951/2020\|EPI_ISL_430646\|2020- | ···················· | ···················· | ···················· | ···················· | ···················· | NN·················· | |
| B | 2836 | hCoV-19/Australia/VIC1329/2020\|EPI_ISL_430696\|2020 | ···················· | ···················· | ···················· | ···················· | ···················· | ···················· | |
| B | 2837 | hCoV-19/Australia/VIC1344/2020\|EPI_ISL_430697\|2020 | ···················· | ···················· | ···················· | ···················· | ···················· | ···················· | |
| B | 2838 | hCoV-19/Australia/VIC1336/2020\|EPI_ISL_430698\|2020 | ···Y················ | ···················· | ···················· | ···················· | NNN················· | ···················· | 1 |
| B | 2839 | hCoV-19/Wales/PHWC-28EF9/2020\|EPI_ISL_431944\|2020- | ···················· | ···················· | ···················· | ···················· | ···················· | ···················· | |
| B | 2840 | hCoV-19/USA/NY-NYUMC433/2020\|EPI_ISL_435500\|2020-0 | ···················· | ···················· | ···················· | ···················· | ···················· | ···················· | |
| B | 2841 | hCoV-19/USA/NY-NYUMC434/2020\|EPI_ISL_435501\|2020-0 | ···················· | ···················· | ···················· | ···················· | ···················· | ···················· | |
| B | 2842 | hCoV-19/USA/NY-NYUMC435/2020\|EPI_ISL_435502\|2020-0 | ···················· | ···················· | ···················· | ···················· | ···················· | ···················· | |
| B | 2843 | hCoV-19/USA/CA-CZB-1009/2020\|EPI_ISL_436650\|2020-0 | ···················· | ···················· | ···················· | ···················· | ···················· | ···················· | |
| B | 2844 | hCoV-19/USA/CA-CZB-1010/2020\|EPI_ISL_436651\|2020-0 | ···················· | ···················· | ···················· | ···················· | ···················· | ···················· | |
| B | 2845 | hCoV-19/USA/MI-MDHHS-SC20113/2020\|EPI_ISL_436800\|2 | ···················· | ···················· | ···················· | ···················· | ···················· | ···················· | |
| B | 2846 | hCoV-19/USA/MI-MDHHS-SC20122/2020\|EPI_ISL_436801\|2 | ···················· | ···················· | ···················· | ···················· | ···················· | ···················· | |
| B | 2847 | hCoV-19/USA/MI-MDHHS-SC20129/2020\|EPI_ISL_436802\|2 | ···················· | ···················· | ···················· | ···················· | ···················· | ···················· | |
| B | 2848 | hCoV-19/USA/MI-MDHHS-SC20162/2020\|EPI_ISL_436803\|2 | ···················· | ···················· | ···················· | ···················· | ···················· | ···················· | |
| B | 2849 | hCoV-19/USA/MI-MDHHS-SC20189/2020\|EPI_ISL_436804\|2 | ···················· | ···················· | ···················· | ···················· | ···················· | ···················· | |
| B | 2850 | hCoV-19/USA/MI-MDHHS-SC20169/2020\|EPI_ISL_436807\|2 | ···················· | ···················· | ···················· | ···················· | ···················· | ···················· | |
| B | 2851 | hCoV-19/USA/MI-MDHHS-SC20179/2020\|EPI_ISL_436808\|2 | ···················· | ···················· | ···················· | ···················· | ···················· | ···················· | |
| B | 2852 | hCoV-19/USA/MI-MDHHS-SC20136/2020\|EPI_ISL_436812\|2 | ···················· | ···················· | ···················· | ···················· | ···················· | ···················· | |
| B | 2853 | hCoV-19/USA/MI-MDHHS-SC20158/2020\|EPI_ISL_436814\|2 | ···················· | ···················· | ···················· | ···················· | ···················· | ···················· | |
| B | 2854 | hCoV-19/USA/MI-MDHHS-SC20130/2020\|EPI_ISL_436815\|2 | ···················· | ···················· | ···················· | ···················· | ···················· | ···················· | |
| B | 2855 | hCoV-19/USA/MI-MDHHS-SC20147/2020\|EPI_ISL_436818\|2 | ···················· | ···················· | ···················· | ···················· | ···················· | ···················· | |
| B | 2856 | hCoV-19/USA/MI-MDHHS-SC20154/2020\|EPI_ISL_436819\|2 | ···················· | ···················· | ···················· | ···················· | ···················· | ···················· | |
| B | 2857 | hCoV-19/USA/MI-MDHHS-SC20139/2020\|EPI_ISL_436820\|2 | ···················· | ···················· | ···················· | ···················· | ···················· | ···················· | |
| B | 2858 | hCoV-19/USA/MI-MDHHS-SC20163/2020\|EPI_ISL_436821\|2 | ···················· | ···················· | ···················· | ···················· | ···················· | ···················· | |
| B | 2859 | hCoV-19/USA/MI-MDHHS-SC20110/2020\|EPI_ISL_436822\|2 | ···················· | ···················· | ···················· | ···················· | ···················· | ···················· | |
| B | 2860 | hCoV-19/USA/MI-MDHHS-SC20161/2020\|EPI_ISL_436823\|2 | ···················· | ···················· | ···················· | ···················· | ···················· | ···················· | |
| B | 2861 | hCoV-19/USA/MI-MDHHS-SC20175/2020\|EPI_ISL_436824\|2 | ···················· | ···················· | ···················· | ···················· | ···················· | ···················· | |
| B | 2862 | hCoV-19/USA/MI-MDHHS-SC20156/2020\|EPI_ISL_436825\|2 | ···················· | ···················· | ···················· | ···················· | ···················· | ···················· | |
| B | 2863 | hCoV-19/USA/MI-MDHHS-SC20171/2020\|EPI_ISL_436827\|2 | ···················· | ···················· | ···················· | ···················· | ···················· | ···················· | |
| B | 2864 | hCoV-19/USA/MI-MDHHS-SC20137/2020\|EPI_ISL_436829\|2 | ···················· | ···················· | ···················· | ···················· | ···················· | ···················· | |
| B | 2865 | hCoV-19/USA/MI-MDHHS-SC20153/2020\|EPI_ISL_436830\|2 | ···················· | ···················· | ···················· | ···················· | ···················· | ···················· | |
| B | 2866 | hCoV-19/USA/MI-MDHHS-SC20107/2020\|EPI_ISL_436831\|2 | ···················· | ···················· | ···················· | ···················· | ···················· | ···················· | |

| | | | | | | | | | |
|---|---|---|---|---|---|---|---|---|---|
| B | 2867 | hCoV-19/USA/MI-MDHHS-SC20132/2020\|EPI_ISL_436833\|2 | ···················· | ···················· | ···················· | ···················· | ···················· | ···················· | |
| B | 2868 | hCoV-19/Austria/CeMM0124/2020\|EPI_ISL_437980\|2020- | ···················· | ···················· | ···················· | ···················· | ···················· | ···················· | |
| B | 2869 | hCoV-19/Austria/CeMM0125/2020\|EPI_ISL_437981\|2020- | ···················· | ···················· | ···················· | ···················· | ···················· | ···················· | |
| B | 2870 | hCoV-19/Austria/CeMM0126/2020\|EPI_ISL_437982\|2020- | ···················· | ···················· | ···················· | ···················· | ···················· | ···················· | |
| B | 2871 | hCoV-19/Scotland/CVR3089/2020\|EPI_ISL_439030\|2020- | ···················· | ···················A | ···················· | ···················· | ···················· | ···················· | 1 |
| B | 2872 | hCoV-19/Scotland/CVR3090/2020\|EPI_ISL_439031\|2020- | ···················· | ···················· | ···················· | ···················· | ···················· | ···················· | |
| B | 2873 | hCoV-19/Scotland/CVR3091/2020\|EPI_ISL_439032\|2020- | ···················· | ···················· | ···················· | ···················· | ···················· | ···················· | |
| B | 2874 | hCoV-19/Scotland/CVR3093/2020\|EPI_ISL_439033\|2020- | ···················· | ···················· | ···················· | ···················· | ···················· | ···················· | |
| B | 2875 | hCoV-19/Scotland/CVR3094/2020\|EPI_ISL_439034\|2020- | ···················· | ···················· | ···················· | ···················· | ···················· | ···················· | |
| B | 2876 | hCoV-19/Scotland/CVR3123/2020\|EPI_ISL_439035\|2020- | ···················· | ···················· | ···················· | ···················· | ···················· | ···················· | |
| B | 2877 | hCoV-19/Scotland/CVR3124/2020\|EPI_ISL_439036\|2020- | ···················· | ···················· | ···················· | ···················· | ···················· | ···················· | |
| B | 2878 | hCoV-19/Scotland/CVR3125/2020\|EPI_ISL_439037\|2020- | ···················· | ···················· | ···················· | ···················· | ···················· | ···················· | |
| B | 2879 | hCoV-19/Scotland/CVR3126/2020\|EPI_ISL_439038\|2020- | ···················· | ···················A | ···················· | ···················· | ···················· | ···················· | 1 |
| B | 2880 | hCoV-19/Scotland/CVR3129/2020\|EPI_ISL_439041\|2020- | ···················· | ···················· | ···················· | ···················· | ···················· | ···················· | |
| B | 2881 | hCoV-19/Scotland/CVR3131/2020\|EPI_ISL_439043\|2020- | ···················· | ···················A | ···················· | ···················· | ···················· | ···················· | 1 |
| B | 2882 | hCoV-19/Scotland/CVR314/2020\|EPI_ISL_439045\|2020-0 | ···················· | ···················· | ···················· | ···················· | ···················· | ···················· | |
| B | 2883 | hCoV-19/Scotland/CVR315/2020\|EPI_ISL_439046\|2020-0 | ···················· | ···················· | ···················· | ···················· | ···················· | ···················· | |
| B | 2884 | hCoV-19/Scotland/CVR318/2020\|EPI_ISL_439048\|2020-0 | ···················· | ···················· | ···················· | ···················· | ···················· | ···················· | |
| B | 2885 | hCoV-19/Scotland/CVR319/2020\|EPI_ISL_439049\|2020-0 | ···················· | ···················· | ···················· | ···················· | ···················· | ···················· | |
| B | 2886 | hCoV-19/Scotland/CVR326/2020\|EPI_ISL_439052\|2020-0 | ···················· | ···················· | ···················· | ···················· | ···················· | ···················· | |
| B | 2887 | hCoV-19/Scotland/CVR337/2020\|EPI_ISL_439056\|2020-0 | ···················· | ···················· | ···················· | ···················· | ···················· | ···················· | |
| B | 2888 | hCoV-19/Scotland/CVR339/2020\|EPI_ISL_439057\|2020-0 | ···················· | ···················· | ···················· | ···················· | ···················· | ···················· | |
| B | 2889 | hCoV-19/Scotland/CVR353/2020\|EPI_ISL_439062\|2020-0 | ···················· | ···················· | ···················· | ···················· | ···················· | ···················· | |
| B | 2890 | hCoV-19/Scotland/CVR355/2020\|EPI_ISL_439063\|2020-0 | ···················· | ···················· | ···················· | ···················· | ···················· | ···················· | |
| B | 2891 | hCoV-19/Scotland/CVR357/2020\|EPI_ISL_439065\|2020-0 | ···················· | ···················· | ···················· | ···················· | ···················· | ···················· | |
| B | 2892 | hCoV-19/Scotland/CVR377/2020\|EPI_ISL_439066\|2020-0 | ···················· | ···················· | ···················· | ···················· | ···················· | ···················· | |
| B | 2893 | hCoV-19/Scotland/CVR378/2020\|EPI_ISL_439067\|2020-0 | ···················· | ···················· | ···················· | ···················· | ···················· | ···················· | |
| B | 2894 | hCoV-19/Scotland/CVR388/2020\|EPI_ISL_439070\|2020-0 | ···················· | ·A·················· | ···················· | ···················· | ···················· | ···················· | |
| B | 2895 | hCoV-19/Scotland/CVR395/2020\|EPI_ISL_439072\|2020-0 | ···················· | ···················· | ···················· | ···················· | ···················· | ···················· | |
| B | 2896 | hCoV-19/Scotland/CVR405/2020\|EPI_ISL_439076\|2020-0 | ···················· | ···················· | ···················· | ···················· | ···················· | ···················· | |
| B | 2897 | hCoV-19/Scotland/CVR453/2020\|EPI_ISL_439080\|2020-0 | ···················· | ···················· | ···················· | ···················· | ···················· | ···················· | |
| B | 2898 | hCoV-19/Scotland/CVR475/2020\|EPI_ISL_439081\|2020-0 | ···················· | ·A·················· | ···················· | ···················· | ···················· | ···················· | |
| B | 2899 | hCoV-19/Scotland/CVR519/2020\|EPI_ISL_439082\|2020-0 | ···················· | ···················· | ···················· | ···················· | ···················· | ···················· | |
| B | 2900 | hCoV-19/Scotland/CVR563/2020\|EPI_ISL_439084\|2020-0 | ···················· | ···················· | ···················· | ···················· | ···················· | ···················· | |
| B | 2901 | hCoV-19/Scotland/CVR565/2020\|EPI_ISL_439085\|2020-0 | ···················· | ···················· | ···················· | ···················· | ···················· | ···················· | |
| B | 2902 | hCoV-19/Scotland/CVR575/2020\|EPI_ISL_439087\|2020-0 | ···················· | ···················· | ···················· | ···················· | ···················· | ···················· | |
| B | 2903 | hCoV-19/Scotland/CVR617/2020\|EPI_ISL_439090\|2020-0 | ···················· | ···················· | ···················· | ···················· | ···················· | ···················· | |
| B | 2904 | hCoV-19/Scotland/CVR620/2020\|EPI_ISL_439091\|2020-0 | ···················· | ···················· | ···················· | ···················· | ···················· | ···················· | |
| B | 2905 | hCoV-19/Scotland/CVR623/2020\|EPI_ISL_439092\|2020-0 | ···················· | ···················· | ···················· | ···················· | ···················· | ···················· | |
| B | 2906 | hCoV-19/Scotland/CVR636/2020\|EPI_ISL_439093\|2020-0 | ···················· | ···················· | ···················· | ···················· | ···················· | ···················· | |
| B | 2907 | hCoV-19/Scotland/CVR660/2020\|EPI_ISL_439095\|2020-0 | ···················· | ···················· | ···················· | ···················· | ···················· | ···················· | |
| B | 2908 | hCoV-19/Scotland/CVR739/2020\|EPI_ISL_439098\|2020-0 | ···················· | ···················· | ···················· | ···················· | ···················· | ···················· | |
| B | 2909 | hCoV-19/England/BRIS-122007/2020\|EPI_ISL_440013\|20 | ···················· | ···················· | ···················· | ···················· | ···················· | ···················· | |
| B | 2910 | hCoV-19/England/CAMB-73642/2020\|EPI_ISL_440014\|202 | ···················· | ···················· | ···················· | ···················· | ···················· | ···················· | |
| B | 2911 | hCoV-19/England/CAMB-72BAE/2020\|EPI_ISL_440017\|202 | ···················· | ···················· | ···················· | ···················· | ···················· | ···················· | |
| B | 2912 | hCoV-19/England/BRIS-122201/2020\|EPI_ISL_440018\|20 | ···················· | ···················· | ···················· | ···················· | ···················· | ···················· | |
| B | 2913 | hCoV-19/England/BRIS-121FFC/2020\|EPI_ISL_440020\|20 | ···················· | ···················· | ···················· | ···················· | ···················· | ···················· | |
| B | 2914 | hCoV-19/England/BRIS-12232C/2020\|EPI_ISL_440021\|20 | ···················· | ···················· | ···················· | ···················· | ···················· | ···················· | |
| B | 2915 | hCoV-19/England/BRIS-122386/2020\|EPI_ISL_440022\|20 | ···················· | ···················· | ···················· | ···················· | ···················· | ···················· | |
| B | 2916 | hCoV-19/England/BRIS-121D3E/2020\|EPI_ISL_440023\|20 | ···················· | ···················· | ···················· | ···················· | ···················· | ···················· | |
| B | 2917 | hCoV-19/England/CAMB-73439/2020\|EPI_ISL_440025\|202 | ···················· | ···················· | ···················· | ···················· | ···················· | ···················· | |
| B | 2918 | hCoV-19/England/BRIS-121F56/2020\|EPI_ISL_440027\|20 | ···················· | ···················· | ···················· | ···················· | ···················· | ···················· | |
| B | 2919 | hCoV-19/England/BRIS-121E3B/2020\|EPI_ISL_440029\|20 | ···················· | ···················· | ···················· | ···················· | ···················· | ···················· | |
| B | 2920 | hCoV-19/England/CAMB-72C8D/2020\|EPI_ISL_440030\|202 | ···················· | ···················· | ···················· | ···················· | ···················· | ···················· | |
| B | 2921 | hCoV-19/England/BRIS-121E86/2020\|EPI_ISL_440032\|20 | ···················· | ···················· | ···················· | ···················· | ···················· | ···················· | |
| B | 2922 | hCoV-19/England/BRIS-1221C8/2020\|EPI_ISL_440033\|20 | ···················· | ···················· | ···················· | ···················· | ···················· | ···················· | |
| B | 2923 | hCoV-19/England/CAMB-72D8A/2020\|EPI_ISL_440034\|202 | ···················· | ···················· | ···················· | ···················· | ···················· | ···················· | |
| B | 2924 | hCoV-19/England/BRIS-121F65/2020\|EPI_ISL_440037\|20 | ···················· | ···················· | ···················· | ···················· | ···················· | ···················· | |
| B | 2925 | hCoV-19/England/BRIS-122025/2020\|EPI_ISL_440038\|20 | ···················· | ···················· | ···················· | ···················· | ···················· | ···················· | |
| B | 2926 | hCoV-19/England/BRIS-121F29/2020\|EPI_ISL_440039\|20 | ···················· | ···················· | ···················· | ···················· | ···················· | ···················· | |
| B | 2927 | hCoV-19/England/CAMB-72C9C/2020\|EPI_ISL_440040\|202 | ·······Y············ | ···················· | ···················· | ···················· | ···················· | ···················· | |
| B | 2928 | hCoV-19/England/CAMB-73457/2020\|EPI_ISL_440041\|202 | ···················· | ···················· | ···················· | ···················· | ···················· | ···················· | |
| B | 2929 | hCoV-19/England/BRIS-122131/2020\|EPI_ISL_440042\|20 | ···················· | ···················· | ···················· | ···················· | ···················· | ···················· | |
| B | 2930 | hCoV-19/England/CAMB-72E0F/2020\|EPI_ISL_440044\|202 | ···················· | ···················· | ···················· | ···················· | ···················· | ···················· | |
| B | 2931 | hCoV-19/England/CAMB-72D99/2020\|EPI_ISL_440045\|202 | ···················· | ···················· | ···················· | ···················· | ···················· | ···················· | |
| B | 2932 | hCoV-19/England/CAMB-72C6F/2020\|EPI_ISL_440046\|202 | ···················· | ···················· | ···················· | ···················· | ···················· | ···················· | |
| B | 2933 | hCoV-19/England/CAMB-736BB/2020\|EPI_ISL_440047\|202 | ···················· | ···················· | ···················· | ···················· | ···················· | ···················· | |
| B | 2934 | hCoV-19/England/BRIS-12223E/2020\|EPI_ISL_440050\|20 | ···················· | ···················· | ···················· | ···················· | ···················· | ···················· | |
| B | 2935 | hCoV-19/England/BRIS-1220AD/2020\|EPI_ISL_440052\|20 | ···················· | ···················· | ···················· | ···················· | ···················· | ···················· | |
| B | 2936 | hCoV-19/England/CAMB-73545/2020\|EPI_ISL_440053\|202 | ···················· | ···················· | ···················· | ···················· | ···················· | ···················· | |
| B | 2937 | hCoV-19/England/BRIS-1220E9/2020\|EPI_ISL_440054\|20 | ···················· | ···················· | ···················· | ···················· | ···················· | ···················· | |
| B | 2938 | hCoV-19/England/CAMB-72EE1/2020\|EPI_ISL_440055\|202 | ···················· | ···················· | ···················· | ···················· | ···················· | ···················· | |
| B | 2939 | hCoV-19/England/BRIS-12234A/2020\|EPI_ISL_440056\|20 | ···················· | ···················· | ···················· | ···················· | ···················· | ···················· | |
| B | 2940 | hCoV-19/England/CAMB-72B53/2020\|EPI_ISL_440057\|202 | ···················· | ···················· | ···················· | ···················· | ···················· | ···················· | |
| B | 2941 | hCoV-19/England/CAMB-72C05/2020\|EPI_ISL_440059\|202 | ···················· | ···················· | ···················· | ···················· | ···················· | ···················· | |
| B | 2942 | hCoV-19/England/CAMB-73554/2020\|EPI_ISL_440060\|202 | ···················· | ···················· | ···················· | ···················· | ···················· | ···················· | |
| B | 2943 | hCoV-19/England/CAMB-72E87/2020\|EPI_ISL_440061\|202 | ···················· | ···················· | ···················· | ···················· | ···················· | ···················· | |
| B | 2944 | hCoV-19/England/CAMB-72D4E/2020\|EPI_ISL_440062\|202 | ···················· | ···················· | ···················· | ···················· | ···················· | ···················· | |

| | | | | | | | | | | |
|---|---|---|---|---|---|---|---|---|---|---|
| B | 2945 | hCoV-19/England/BRIS-12225C/2020\|EPI_ISL_440063\|20 | .................... | .................... | .................... | .................... | .................... | .................... | | |
| B | 2946 | hCoV-19/England/CAMB-72B44/2020\|EPI_ISL_440064\|202 | .................... | .................... | .................... | .................... | .................... | .................... | | |
| B | 2947 | hCoV-19/England/BRIS-122140/2020\|EPI_ISL_440065\|20 | .................... | .................... | .................... | .................... | .................... | .................... | | |
| B | 2948 | hCoV-19/England/CAMB-72DF3/2020\|EPI_ISL_440066\|202 | .................... | .................... | .................... | .................... | .................... | .................... | | |
| B | 2949 | hCoV-19/England/BRIS-12231D/2020\|EPI_ISL_440068\|20 | .................... | .................... | .................... | .................... | .................... | .................... | | |
| B | 2950 | hCoV-19/England/CAMB-72F1B/2020\|EPI_ISL_440069\|202 | .................... | .................... | .................... | .................... | .................... | .................... | | |
| B | 2951 | hCoV-19/England/BRIS-121E4A/2020\|EPI_ISL_440070\|20 | .................... | .................... | .................... | .................... | .................... | .................... | | |
| B | 2952 | hCoV-19/England/BRIS-121E59/2020\|EPI_ISL_440071\|20 | .................... | .................... | .................... | .................... | .................... | .................... | | |
| B | 2953 | hCoV-19/England/CAMB-72D11/2020\|EPI_ISL_440072\|202 | .................... | .................... | .................... | .................... | .................... | .................... | | |
| B | 2954 | hCoV-19/England/CAMB-735CD/2020\|EPI_ISL_440073\|202 | .................... | .................... | .................... | .................... | .................... | .................... | | |
| B | 2955 | hCoV-19/England/CAMB-72EA5/2020\|EPI_ISL_440074\|202 | .................... | .................... | .................... | .................... | .................... | .................... | | |
| B | 2956 | hCoV-19/England/CAMB-72E5A/2020\|EPI_ISL_440075\|202 | .................... | .................... | .................... | .................... | .................... | .................... | | |
| B | 2957 | hCoV-19/England/CAMB-72B80/2020\|EPI_ISL_440076\|202 | .................... | .................... | .................... | .................... | .................... | .................... | | |
| B | 2958 | hCoV-19/England/CAMB-72C14/2020\|EPI_ISL_440077\|202 | .................... | .................... | .................... | .................... | .................... | .................... | | |
| B | 2959 | hCoV-19/England/CAMB-72D3F/2020\|EPI_ISL_440079\|202 | .................... | .................... | .................... | .................... | .................... | .................... | | |
| B | 2960 | hCoV-19/England/CAMB-72EB4/2020\|EPI_ISL_440080\|202 | .................... | .................... | .................... | .................... | .................... | .................... | | |
| B | 2961 | hCoV-19/England/BRIS-12216E/2020\|EPI_ISL_440081\|20 | .................... | .................... | .................... | .................... | .................... | .................... | | |
| B | 2962 | hCoV-19/England/CAMB-735BE/2020\|EPI_ISL_440085\|202 | .................... | .................... | .................... | .................... | .................... | .................... | | |
| B | 2963 | hCoV-19/England/CAMB-73466/2020\|EPI_ISL_440086\|202 | .................... | .............A...... | .................... | .................... | .................... | .................... | | |
| B | 2964 | hCoV-19/England/BRIS-1220CB/2020\|EPI_ISL_440087\|20 | .................... | .................... | .................... | .................... | .................... | .................... | | |
| B | 2965 | hCoV-19/England/CAMB-72E4B/2020\|EPI_ISL_440088\|202 | .................... | .................... | .................... | .................... | .................... | .................... | | |
| B | 2966 | hCoV-19/England/CAMB-72B9F/2020\|EPI_ISL_440090\|202 | .................... | .................... | .................... | .................... | .................... | .................... | | |
| B | 2967 | hCoV-19/England/BRIS-1221D7/2020\|EPI_ISL_440091\|20 | .................... | .................... | .................... | .................... | .................... | .................... | | |
| B | 2968 | hCoV-19/England/BRIS-121D10/2020\|EPI_ISL_440092\|20 | .................... | ...................A | .................... | .................... | .................... | .................... | 1 | |
| B | 2969 | hCoV-19/England/BRIS-121EA4/2020\|EPI_ISL_440093\|20 | .................... | .................... | .................... | .................... | .................... | .................... | | |
| B | 2970 | hCoV-19/England/BRIS-12233B/2020\|EPI_ISL_440094\|20 | .................... | .................... | .................... | .................... | .................... | .................... | | |
| B | 2971 | hCoV-19/England/BRIS-121E77/2020\|EPI_ISL_440095\|20 | .................... | .................... | .................... | .................... | .................... | .................... | | |
| B | 2972 | hCoV-19/England/CAMB-72B62/2020\|EPI_ISL_440096\|202 | .................... | .................... | .................... | .................... | .................... | .................... | | |
| B | 2973 | hCoV-19/England/CAMB-72F2A/2020\|EPI_ISL_440097\|202 | .................... | .................... | .................... | .................... | .................... | .................... | | |
| B | 2974 | hCoV-19/England/BRIS-121DB6/2020\|EPI_ISL_440098\|20 | .................... | .............A...... | .................... | .................... | .................... | .................... | | |
| B | 2975 | hCoV-19/England/CAMB-72CF6/2020\|EPI_ISL_440099\|202 | .................... | .................... | .................... | .................... | .................... | .................... | | |
| B | 2976 | hCoV-19/England/CAMB-7A99D/2020\|EPI_ISL_441344\|202 | .................... | .................... | .................... | .................... | .................... | .................... | | |
| B | 2977 | hCoV-19/England/CAMB-7A836/2020\|EPI_ISL_441345\|202 | .................... | .................... | .................... | .................... | .................... | .................... | | |
| B | 2978 | hCoV-19/England/CAMB-7A0A7/2020\|EPI_ISL_441346\|202 | .................... | .................... | .................... | .................... | .................... | .................... | | |
| B | 2979 | hCoV-19/England/CAMB-7A687/2020\|EPI_ISL_441347\|202 | .................... | .................... | .................... | .................... | .................... | .................... | | |
| B | 2980 | hCoV-19/England/CAMB-78DDF/2020\|EPI_ISL_441349\|202 | .................... | .................... | .................... | .................... | .................... | .................... | | |
| B | 2981 | hCoV-19/Northern Ireland/NIRE-101EF5/2020\|EPI_ISL_ | .................... | .................... | .................... | .................... | .................... | .................... | | |
| B | 2983 | hCoV-19/Northern Ireland/NIRE-101F3E/2020\|EPI_ISL_ | .................... | .................... | .................... | .................... | .................... | .................... | | |
| B | 2985 | hCoV-19/Northern Ireland/NIRE-101F6B/2020\|EPI_ISL_ | .................... | .................... | .................... | .................... | .................... | .................... | | |
| B | 2986 | hCoV-19/Northern Ireland/NIRE-101F7A/2020\|EPI_ISL_ | .................... | .................... | .................... | .................... | .................... | .................... | | |
| B | 2988 | hCoV-19/Northern Ireland/NIRE-101FD4/2020\|EPI_ISL_ | .................... | .................... | .................... | .................... | .................... | .................... | | |
| B | 2989 | hCoV-19/Northern Ireland/NIRE-101FE3/2020\|EPI_ISL_ | .................... | .................... | .................... | .................... | .NN................. | .................... | | |
| B | 2992 | hCoV-19/Northern Ireland/NIRE-10201C/2020\|EPI_ISL_ | .................... | .................... | .................... | .................... | .................... | .................... | | |
| B | 2994 | hCoV-19/Northern Ireland/NIRE-10203A/2020\|EPI_ISL_ | .................... | .................... | .................... | .................... | .................... | .................... | | |
| B | 2995 | hCoV-19/Northern Ireland/NIRE-102049/2020\|EPI_ISL_ | .................... | .................... | .................... | .................... | .................... | .................... | | |
| B | 2996 | hCoV-19/Northern Ireland/NIRE-102058/2020\|EPI_ISL_ | .................... | .................... | .................... | .................... | .................... | .................... | | |
| B | 2997 | hCoV-19/Northern Ireland/NIRE-102067/2020\|EPI_ISL_ | .................... | .................... | .................... | .................... | .................... | .................... | | |
| B | 2998 | hCoV-19/Northern Ireland/NIRE-102076/2020\|EPI_ISL_ | .................... | .................... | .................... | .................... | .................... | .................... | | |
| B | 2999 | hCoV-19/Northern Ireland/NIRE-102094/2020\|EPI_ISL_ | .................... | .................... | .................... | .................... | .................... | .................... | | |
| B | 3001 | hCoV-19/Northern Ireland/NIRE-1020C1/2020\|EPI_ISL_ | .................... | .................... | .................... | .................... | .................... | .................... | | |
| B | 3004 | hCoV-19/Northern Ireland/NIRE-1020FE/2020\|EPI_ISL_ | .................... | .................... | .................... | .................... | .................... | .................... | | |
| B | 3006 | hCoV-19/Northern Ireland/NIRE-1022BC/2020\|EPI_ISL_ | .................... | .................... | .................... | .................... | .NNN................ | .................... | | 1 |
| B | 3008 | hCoV-19/Northern Ireland/NIRE-1022DA/2020\|EPI_ISL_ | .................... | .................... | .................... | .................... | .................... | .................... | | |
| B | 3009 | hCoV-19/England/CAMB-7E9A8/2020\|EPI_ISL_442610\|202 | .................... | .................... | .................... | .................... | .................... | .................... | | |
| B | 3010 | hCoV-19/England/CAMB-7E93F/2020\|EPI_ISL_442611\|202 | .................... | .................... | .................... | .................... | .................... | .................... | | |
| B | 3011 | hCoV-19/England/CAMB-8173F/2020\|EPI_ISL_442613\|202 | .................... | .................... | .................... | .................... | .................... | .................... | | |
| B | 3012 | hCoV-19/England/CAMB-7E8C9/2020\|EPI_ISL_442614\|202 | .................... | .................... | .................... | .................... | .................... | .................... | | |
| B | 3013 | hCoV-19/England/CAMB-7E8AB/2020\|EPI_ISL_442615\|202 | .................... | .................... | .................... | .................... | .................... | .................... | | |
| B | 3014 | hCoV-19/England/CAMB-7EA5A/2020\|EPI_ISL_442616\|202 | .................... | .................... | .................... | .................... | .................... | .................... | | |
| B | 3015 | hCoV-19/England/CAMB-8167E/2020\|EPI_ISL_442618\|202 | .................... | .................... | .................... | .................... | .................... | .................... | | |
| B | 3016 | hCoV-19/England/CAMB-81993/2020\|EPI_ISL_442620\|202 | .................... | .................... | .................... | .................... | .................... | .................... | | |
| B | 3017 | hCoV-19/England/CAMB-80958/2020\|EPI_ISL_442622\|202 | .................... | .................... | .................... | .................... | .................... | .................... | | |
| B | 3018 | hCoV-19/England/CAMB-814FC/2020\|EPI_ISL_442623\|202 | .................... | .................... | .................... | .................... | .................... | .................... | | |
| B | 3019 | hCoV-19/England/BRIS-1238FF/2020\|EPI_ISL_442624\|20 | .................... | .................... | .................... | .................... | .................... | .................... | | |
| B | 3020 | hCoV-19/England/BRIS-12321F/2020\|EPI_ISL_442625\|20 | .................... | .............A...... | .................... | .................... | .................... | .................... | | |
| B | 3021 | hCoV-19/England/BRIS-123507/2020\|EPI_ISL_442626\|20 | .................... | .................... | .................... | .................... | .................... | .................... | | |
| B | 3022 | hCoV-19/England/BRIS-123789/2020\|EPI_ISL_442627\|20 | .................... | .................... | .................... | .................... | .................... | .................... | | |
| B | 3023 | hCoV-19/England/BRIS-123015/2020\|EPI_ISL_442628\|20 | .................... | .................... | .................... | .................... | .................... | .................... | | |
| B | 3024 | hCoV-19/England/BRIS-1231D6/2020\|EPI_ISL_442629\|20 | .................... | ....A............... | .................... | .................... | .................... | .................... | | |
| B | 3025 | hCoV-19/England/BRIS-1232A6/2020\|EPI_ISL_442630\|20 | .................... | .................... | .................... | .................... | .................... | .................... | | |
| B | 3026 | hCoV-19/England/BRIS-123367/2020\|EPI_ISL_442675\|20 | .................... | .................... | .................... | .................... | .................... | .................... | | |
| B | 3027 | hCoV-19/England/BRIS-12315E/2020\|EPI_ISL_442676\|20 | .................... | .................... | .................... | .................... | .................... | .................... | | |
| B | 3028 | hCoV-19/England/BRIS-123455/2020\|EPI_ISL_442679\|20 | .................... | .................... | .................... | .................... | .................... | .................... | | |
| B | 3029 | hCoV-19/England/BRIS-1230F7/2020\|EPI_ISL_442680\|20 | .................... | .................... | .................... | .................... | .................... | .................... | | |
| B | 3030 | hCoV-19/England/BRIS-1235F8/2020\|EPI_ISL_442681\|20 | .................... | .................... | .................... | .................... | .................... | .................... | | |
| B | 3031 | hCoV-19/England/BRIS-12380E/2020\|EPI_ISL_442682\|20 | .................... | .................... | .................... | .................... | .................... | .................... | | |
| B | 3032 | hCoV-19/England/BRIS-123033/2020\|EPI_ISL_442683\|20 | .................... | .................... | .................... | .................... | .................... | .................... | | |
| B | 3033 | hCoV-19/England/BRIS-123ACC/2020\|EPI_ISL_442684\|20 | .................... | .................... | .................... | .................... | .................... | .................... | | |

| | | | | | | | |
|---|---|---|---|---|---|---|---|
| B | 3034 hCoV-19/England/BRIS-123C7B/2020\|EPI_ISL_442685\|20 | ···················· | ···················· | ···················· | ···················· | ···················· | ···················· |
| B | 3035 hCoV-19/England/BRIS-12330D/2020\|EPI_ISL_442686\|20 | ···················· | ···················· | ···················· | ···················· | ···················· | ···················· |
| B | 3036 hCoV-19/England/BRIS-123B9C/2020\|EPI_ISL_442688\|20 | ···················· | ···················· | ···················· | ···················· | ···················· | ···················· |
| B | 3037 hCoV-19/England/BRIS-12375C/2020\|EPI_ISL_442689\|20 | ···················· | ···················· | ···················· | ···················· | ···················· | ···················· |
| B | 3038 hCoV-19/England/BRIS-1232D3/2020\|EPI_ISL_442690\|20 | ···················· | ···················· | ···················· | ···················· | ···················· | ···················· |
| B | 3039 hCoV-19/England/BRIS-12368C/2020\|EPI_ISL_442691\|20 | ···················· | ···················· | ···················· | ···················· | ···················· | ···················· |
| B | 3040 hCoV-19/England/BRIS-1237D4/2020\|EPI_ISL_442692\|20 | ···················· | ···················· | ···················· | ···················· | ···················· | ···················· |
| B | 3041 hCoV-19/England/BRIS-123798/2020\|EPI_ISL_442694\|20 | ···················· | ·A·················· | ···················· | ···················· | ···················· | ···················· |
| B | 3042 hCoV-19/England/BRIS-1239A1/2020\|EPI_ISL_442695\|20 | ···················· | ···················· | ···················· | ···················· | ···················· | ···················· |
| B | 3043 hCoV-19/England/BRIS-123376/2020\|EPI_ISL_442696\|20 | ···················· | ···················· | ···················· | ···················· | ···················· | ···················· |
| B | 3044 hCoV-19/England/BRIS-12374D/2020\|EPI_ISL_442697\|20 | ···················· | ···················· | ···················· | ···················· | ···················· | ···················· |
| B | 3045 hCoV-19/England/BRIS-123B32/2020\|EPI_ISL_442699\|20 | ···················· | ···················· | ···················· | ···················· | ···················· | ···················· |
| B | 3046 hCoV-19/England/BRIS-1259FA/2020\|EPI_ISL_443928\|20 | ···················· | ···················· | ···················· | ···················· | ···················· | ···················· |
| B | 3047 hCoV-19/England/BRIS-125DA3/2020\|EPI_ISL_443929\|20 | ···················· | ···················· | ···················· | ···················· | ···················· | ···················· |
| B | 3048 hCoV-19/England/BRIS-124946/2020\|EPI_ISL_443940\|20 | ···················· | ···················· | ···················· | ···················· | ···················· | ···················· |
| B | 3049 hCoV-19/England/BRIS-124F26/2020\|EPI_ISL_443941\|20 | ···················· | ···················· | ···················· | ···················· | ···················· | ···················· |
| B | 3050 hCoV-19/England/BRIS-124B22/2020\|EPI_ISL_443942\|20 | ···················· | ···················· | ···················· | ···················· | ···················· | ···················· |
| B | 3051 hCoV-19/England/BRIS-1253DE/2020\|EPI_ISL_443943\|20 | ···················· | ···················· | ···················· | ···················· | ···················· | ···················· |
| B | 3052 hCoV-19/England/BRIS-124EB0/2020\|EPI_ISL_443944\|20 | ···················· | ···················· | ···················· | ···················· | ···················· | ···················· |
| B | 3053 hCoV-19/England/BRIS-124991/2020\|EPI_ISL_443945\|20 | ···················· | ···················· | ···················· | ···················· | ···················· | ···················· |
| B | 3054 hCoV-19/England/BRIS-124D77/2020\|EPI_ISL_443946\|20 | ···················· | ···················· | ···················· | ···················· | ···················· | ···················· |
| B | 3055 hCoV-19/England/BRIS-1252FF/2020\|EPI_ISL_443947\|20 | ···················· | ···················· | ···················· | ···················· | ···················· | ···················· |
| B | 3056 hCoV-19/England/BRIS-125E19/2020\|EPI_ISL_443948\|20 | ···················· | ···················· | ···················· | ···················· | ···················· | ···················· |
| B | 3057 hCoV-19/England/BRIS-125CF1/2020\|EPI_ISL_443949\|20 | ···················· | ···················· | ···················· | ···················· | ···················· | ···················· |
| B | 3058 hCoV-19/England/BRIS-124BB9/2020\|EPI_ISL_443950\|20 | ···················· | ···················· | ···················· | ···················· | ···················· | ···················· |
| B | 3059 hCoV-19/England/BRIS-125893/2020\|EPI_ISL_443951\|20 | ···················· | ···················· | ···················· | ···················· | ···················· | ···················· |
| B | 3060 hCoV-19/England/BRIS-12507D/2020\|EPI_ISL_443952\|20 | ···················· | ···················· | ···················· | ···················· | ···················· | ···················· |
| B | 3061 hCoV-19/England/BRIS-125927/2020\|EPI_ISL_443953\|20 | ···················· | ···················· | ···················· | ···················· | ···················· | ···················· |
| B | 3062 hCoV-19/England/BRIS-124DB3/2020\|EPI_ISL_443954\|20 | ···················· | ···················· | ···················· | ···················· | ···················· | ···················· |
| B | 3063 hCoV-19/England/BRIS-125E82/2020\|EPI_ISL_443955\|20 | ···················· | ···················· | ···················· | ···················· | ···················· | ···················· |
| B | 3064 hCoV-19/England/BRIS-125DEF/2020\|EPI_ISL_443956\|20 | ···················· | ···················· | ···················· | ···················· | ···················· | ···················· |
| B | 3065 hCoV-19/England/BRIS-125408/2020\|EPI_ISL_443957\|20 | ···················· | ···················· | ···················· | ···················· | ···················· | ···················· |
| B | 3066 hCoV-19/England/BRIS-125189/2020\|EPI_ISL_443958\|20 | ···················· | ···················· | ···················· | ···················· | ···················· | ···················· |
| B | 3067 hCoV-19/England/BRIS-1258A2/2020\|EPI_ISL_443959\|20 | ···················· | ···················· | ···················· | ···················· | ···················· | ···················· |
| B | 3068 hCoV-19/England/BRIS-125ACA/2020\|EPI_ISL_443961\|20 | ···················· | ···················· | ···················· | ···················· | ···················· | ···················· |
| B | 3069 hCoV-19/USA/WA-QDX-02-2/2020\|EPI_ISL_454692\|2020-0 | ···················· | ···················· | ···················· | ···················· | ···················· | ···················· |
| B | 3070 hCoV-19/USA/CA-QDX-03/2020\|EPI_ISL_454693\|2020-03- | ···················· | ···················· | ···················· | ···················· | ···················· | ···················· |
| B | 3071 hCoV-19/Henan/HN01/2020\|EPI_ISL_605929\|2020-02-24 | ···················· | ···················· | ···················· | ···················· | ···················· | ···················· |
| B | 3072 hCoV-19/Henan/HN04/2020\|EPI_ISL_605930\|2020-02-24 | ···················· | ···················· | ···················· | ···················· | ···················· | ···················· |
| B | 3076 hCoV-19/Australia/VIC964/2020\|EPI_ISL_430655\|2020- | ···················· | ···················· | ···················· | ···················· | ···················· | ···················· |
| B | 3082 hCoV-19/Australia/VIC1067/2020\|EPI_ISL_430670\|2020 | ···················· | ···················· | ···················· | ···················· | ···················· | ···················· |
| B | 3084 hCoV-19/Australia/VIC1178/2020\|EPI_ISL_430674\|2020 | ···················· | ···················· | ···················· | ···················· | ···················· | ·····R·············· |
| B | 3088 hCoV-19/Australia/VIC1301/2020\|EPI_ISL_430679\|2020 | ···················· | ···················· | ···················· | ···················· | ···················· | ···················· |
| B | 3089 hCoV-19/Australia/VIC1302/2020\|EPI_ISL_430680\|2020 | ···················· | ···················· | ···················· | ···················· | ···················· | ···················· |
| B | 3090 hCoV-19/Australia/VIC1314/2020\|EPI_ISL_430681\|2020 | ···················· | ···················· | ···················· | ···················· | ···················· | ···················· |
| B | 3091 hCoV-19/Australia/VIC1334/2020\|EPI_ISL_430682\|2020 | ···················· | ···················· | ···················· | ···················· | ···················· | ···················· |
| B | 3092 hCoV-19/Australia/VIC1342/2020\|EPI_ISL_430683\|2020 | ···················· | ···················· | ···················· | ···················· | ···················· | ···················· |
| B | 3093 hCoV-19/Australia/VIC1343/2020\|EPI_ISL_430684\|2020 | ···················· | ···················· | ···················· | ···················· | ···················· | ···················· |
| B | 3095 hCoV-19/Australia/VIC1275/2020\|EPI_ISL_430687\|2020 | ···················· | ···················· | ···················· | ···················· | ···················· | ···················· |
| B | 3096 hCoV-19/Australia/VIC1315/2020\|EPI_ISL_430689\|2020 | ···················· | ···················· | ···················· | ···················· | ···················· | ···················· |
| B | 3097 hCoV-19/Australia/VIC1316/2020\|EPI_ISL_430690\|2020 | ···················· | ···················· | ···················· | ···················· | ···················· | ···················· |
| B | 3099 hCoV-19/Australia/VIC1318/2020\|EPI_ISL_430693\|2020 | ···················· | ···················· | ···················· | ···················· | ···················· | ···················· |
| B | 3100 hCoV-19/Australia/VIC1303/2020\|EPI_ISL_430694\|2020 | ···················· | ···················· | ···················· | ···················· | ···················· | ···················· |
| B | 3102 hCoV-19/USA/WA-S379/2020\|EPI_ISL_434139\|2020-03-27 | ···················· | ···················· | ···················· | ···················· | ···················· | ···················· |
| B | 3103 hCoV-19/USA/WA-S380/2020\|EPI_ISL_434140\|2020-03-27 | ···················· | ···················· | ···················· | ···················· | ···················· | ···················· |
| B | 3104 hCoV-19/USA/WA-S381/2020\|EPI_ISL_434141\|2020-03-29 | ···················· | ···················· | ···················· | ···················· | ···················· | ···················· |
| B | 3105 hCoV-19/USA/WA-S382/2020\|EPI_ISL_434142\|2020-03-27 | ···················· | ···················· | ···················· | ···················· | ···················· | ···················· |
| B | 3106 hCoV-19/USA/WA-S383/2020\|EPI_ISL_434143\|2020-03-27 | ···················· | ···················· | ···················· | ···················· | ···················· | ···················· |
| B | 3107 hCoV-19/USA/WA-S384/2020\|EPI_ISL_434144\|2020-03-27 | ···················· | ···················· | ···················· | ···················· | ···················· | ···················· |
| B | 3108 hCoV-19/USA/WA-S385/2020\|EPI_ISL_434145\|2020-03-28 | ···················· | ···················· | ···················· | ···················· | ···················· | ···················· |
| B | 3109 hCoV-19/USA/WA-S386/2020\|EPI_ISL_434146\|2020-03-26 | ···················· | ···················· | ···················· | ···················· | ···················· | ···················· |
| B | 3110 hCoV-19/USA/WA-S387/2020\|EPI_ISL_434147\|2020-03-27 | ···················· | ···················· | ···················· | ···················· | ···················· | ···················· |
| B | 3111 hCoV-19/USA/WA-S388/2020\|EPI_ISL_434148\|2020-03-27 | ···················· | ···················· | ···················· | ···················· | ···················· | ···················· |
| B | 3112 hCoV-19/USA/WA-S389/2020\|EPI_ISL_434149\|2020-03-28 | ···················· | ···················· | ···················· | ···················· | ···················· | ···················· |
| B | 3113 hCoV-19/USA/WA-S390/2020\|EPI_ISL_434150\|2020-03-27 | ···················· | ···················· | ···················· | ···················· | ···················· | ···················· |
| B | 3114 hCoV-19/USA/WA-S392/2020\|EPI_ISL_434152\|2020-03-30 | ···················· | ···················· | ···················· | ···················· | ···················· | ···················· |
| B | 3115 hCoV-19/USA/WA-S393/2020\|EPI_ISL_434153\|2020-03-28 | ···················· | ···················· | ···················· | ···················· | ···················· | ···················· |
| B | 3116 hCoV-19/USA/WA-S394/2020\|EPI_ISL_434154\|2020-03-30 | ···················· | ···················· | ···················· | ···················· | ···················· | ···················· |
| B | 3117 hCoV-19/USA/WA-S395/2020\|EPI_ISL_434155\|2020-03-30 | ···················· | ···················· | ···················· | ···················· | ···················· | ···················· |
| B | 3118 hCoV-19/USA/WA-S396/2020\|EPI_ISL_434156\|2020-03-25 | ···················· | ···················· | ···················· | ···················· | ···················· | ···················· |
| B | 3119 hCoV-19/USA/WA-S397/2020\|EPI_ISL_434157\|2020-03-25 | ···················· | ···················· | ···················· | ···················· | ···················· | ···················· |
| B | 3120 hCoV-19/USA/WA-S398/2020\|EPI_ISL_434158\|2020-03-30 | ···················· | ···················· | ···················· | ···················· | ···················· | ···················· |
| B | 3121 hCoV-19/USA/WA-S399/2020\|EPI_ISL_434159\|2020-03-28 | ···················· | ···················· | ···················· | ···················· | ···················· | ···················· |
| B | 3122 hCoV-19/USA/WA-S400/2020\|EPI_ISL_434160\|2020-03-28 | ···················· | ···················· | ···················· | ···················· | ···················· | ···················· |
| B | 3123 hCoV-19/USA/WA-S401/2020\|EPI_ISL_434161\|2020-03-28 | ···················· | ···················· | ···················· | ···················· | ···················· | ···················· |
| B | 3124 hCoV-19/USA/WA-S402/2020\|EPI_ISL_434162\|2020-03-28 | ···················· | ···················· | ···················· | ···················· | ···················· | ···················· |
| B | 3125 hCoV-19/USA/WA-S403/2020\|EPI_ISL_434163\|2020-03-26 | ···················· | ···················· | ···················· | ···················· | ···················· | ···················· |
| B | 3126 hCoV-19/USA/WA-S404/2020\|EPI_ISL_434164\|2020-03-26 | ···················· | ···················· | ···················· | ···················· | ···················· | ···················· |

| | | | | | | | | |
|---|---|---|---|---|---|---|---|---|
| B | 3127 hCoV-19/USA/WA-S405/2020\|EPI_ISL_434165\|2020-03-25 | .................... | .................... | .................... | .................... | .................... | .................... | |
| B | 3128 hCoV-19/USA/WA-S406/2020\|EPI_ISL_434166\|2020-03-26 | .................... | .................... | .................... | .................... | .................... | .................... | |
| B | 3129 hCoV-19/USA/WA-S407/2020\|EPI_ISL_434167\|2020-03-28 | .................... | .................... | .................... | .................... | .................... | .................... | |
| B | 3130 hCoV-19/USA/WA-S409/2020\|EPI_ISL_434169\|2020-03-28 | .................... | .................... | .................... | .................... | .................... | .................... | |
| B | 3131 hCoV-19/USA/WA-S410/2020\|EPI_ISL_434170\|2020-03-29 | .................... | .................... | .................... | .................... | .................... | .................... | |
| B | 3132 hCoV-19/USA/WA-S411/2020\|EPI_ISL_434171\|2020-03-28 | .................... | .................... | .................... | .................... | .................... | .................... | |
| B | 3133 hCoV-19/USA/WA-S412/2020\|EPI_ISL_434172\|2020-03-30 | .................... | .................... | .................... | .................... | .................... | .................... | |
| B | 3134 hCoV-19/USA/WA-S413/2020\|EPI_ISL_434173\|2020-03-26 | .................... | .................... | .................... | .................... | .................... | .................... | |
| B | 3135 hCoV-19/USA/WA-S415/2020\|EPI_ISL_434175\|2020-03-28 | .................... | .................... | .................... | .................... | .................... | .................... | |
| B | 3136 hCoV-19/USA/WA-S416/2020\|EPI_ISL_434176\|2020-03-29 | .................... | .................... | .................... | .................... | .................... | .................... | |
| B | 3137 hCoV-19/USA/WA-S417/2020\|EPI_ISL_434177\|2020-03-28 | .................... | .................... | .................... | .................... | .................... | .................... | |
| B | 3138 hCoV-19/USA/WA-S418/2020\|EPI_ISL_434178\|2020-03-25 | .................... | .................... | .................... | .................... | .................... | .................... | |
| B | 3139 hCoV-19/USA/WA-S419/2020\|EPI_ISL_434179\|2020-03-28 | .................... | .................... | .................... | .................... | .................... | .................... | |
| B | 3140 hCoV-19/USA/LA-SR0169/2020\|EPI_ISL_435470\|2020-04- | .................... | .................... | .................... | .................... | .................... | .................... | |
| B | 3141 hCoV-19/USA/LA-SR0170/2020\|EPI_ISL_435471\|2020-04- | .................... | .................... | .................... | .................... | .................... | .................... | |
| B | 3142 hCoV-19/USA/CA-SR0175/2020\|EPI_ISL_435473\|2020-04- | .................... | .................... | .................... | .................... | .................... | .................... | |
| B | 3143 hCoV-19/USA/CA-SR0176/2020\|EPI_ISL_435474\|2020-04- | .................... | .A.................. | .................... | .................... | .................... | .................... | |
| B | 3144 hCoV-19/USA/NY-NYUMC408/2020\|EPI_ISL_435475\|2020-0 | .................... | .................... | .................... | .................... | .................... | .................... | |
| B | 3145 hCoV-19/USA/NY-NYUMC409/2020\|EPI_ISL_435476\|2020-0 | .................... | .................... | .................... | .................... | .................... | .................... | |
| B | 3146 hCoV-19/USA/NY-NYUMC410/2020\|EPI_ISL_435477\|2020-0 | .................... | .................... | .................... | .................... | .................... | .................... | |
| B | 3147 hCoV-19/USA/NY-NYUMC411/2020\|EPI_ISL_435478\|2020-0 | .................... | .................... | .................... | .................... | .................... | .................... | |
| B | 3148 hCoV-19/USA/NY-NYUMC412/2020\|EPI_ISL_435479\|2020-0 | .................... | .................... | .................... | .................... | .................... | .................... | |
| B | 3149 hCoV-19/USA/NY-NYUMC413/2020\|EPI_ISL_435480\|2020-0 | .................... | .................... | .................... | .................... | .................... | .................... | |
| B | 3150 hCoV-19/USA/NY-NYUMC414/2020\|EPI_ISL_435481\|2020-0 | .................... | .................... | .................... | .................... | .................... | .................... | |
| B | 3151 hCoV-19/USA/NY-NYUMC415/2020\|EPI_ISL_435482\|2020-0 | .................... | .................... | .................... | .................... | .................... | .................... | |
| B | 3152 hCoV-19/USA/NJ-NYUMC416/2020\|EPI_ISL_435483\|2020-0 | .................... | .................... | .................... | .................... | .................... | .................... | |
| B | 3153 hCoV-19/USA/NY-NYUMC417/2020\|EPI_ISL_435484\|2020-0 | .................... | .................... | .................... | .................... | .................... | .................... | |
| B | 3154 hCoV-19/USA/NY-NYUMC418/2020\|EPI_ISL_435485\|2020-0 | .................... | .................... | .................... | .................... | .................... | .................... | |
| B | 3155 hCoV-19/USA/NY-NYUMC419/2020\|EPI_ISL_435486\|2020-0 | .................... | .................... | .................... | .................... | .................... | .................... | |
| B | 3156 hCoV-19/USA/NY-NYUMC420/2020\|EPI_ISL_435487\|2020-0 | .................... | .................... | .................... | .................... | .................... | .................... | |
| B | 3157 hCoV-19/USA/NJ-NYUMC421/2020\|EPI_ISL_435488\|2020-0 | .................... | .................... | .................... | .................... | .................... | .................... | |
| B | 3158 hCoV-19/USA/NY-NYUMC422/2020\|EPI_ISL_435489\|2020-0 | .................... | .................... | .................... | .................... | .................... | .................... | |
| B | 3159 hCoV-19/USA/NY-NYUMC423/2020\|EPI_ISL_435490\|2020-0 | .................... | .................... | .................... | .................... | .................... | .................... | |
| B | 3160 hCoV-19/USA/NY-NYUMC424/2020\|EPI_ISL_435491\|2020-0 | .................... | .................... | .................... | .................... | .................... | .................... | |
| B | 3161 hCoV-19/USA/NY-NYUMC425/2020\|EPI_ISL_435492\|2020-0 | .................... | .................... | .................... | .................... | .................... | .................... | |
| B | 3162 hCoV-19/USA/NY-NYUMC426/2020\|EPI_ISL_435493\|2020-0 | .................... | .................... | .................... | .................... | .................... | .................... | |
| B | 3163 hCoV-19/USA/NY-NYUMC427/2020\|EPI_ISL_435494\|2020-0 | .................... | .................... | .................... | .................... | .................... | .................... | |
| B | 3164 hCoV-19/USA/NY-NYUMC428/2020\|EPI_ISL_435495\|2020-0 | .................... | .................... | .................... | .................... | .................... | .................... | |
| B | 3165 hCoV-19/USA/NY-NYUMC429/2020\|EPI_ISL_435496\|2020-0 | .................... | .................... | .................... | .................... | .................... | .................... | |
| B | 3166 hCoV-19/USA/NY-NYUMC430/2020\|EPI_ISL_435497\|2020-0 | .................... | .................... | .................... | .................... | .................... | .................... | |
| B | 3167 hCoV-19/USA/NY-NYUMC431/2020\|EPI_ISL_435498\|2020-0 | .................... | .................... | .................... | .................... | .................... | .................... | |
| B | 3168 hCoV-19/USA/NY-NYUMC432/2020\|EPI_ISL_435499\|2020-0 | .................... | .................... | .................... | .................... | .................... | .................... | |
| B | 3169 hCoV-19/Scotland/CVR2939/2020\|EPI_ISL_439000\|2020- | .................... | .................... | .................... | .................... | .................... | .................... | |
| B | 3170 hCoV-19/Scotland/CVR2987/2020\|EPI_ISL_439003\|2020- | .................... | ...................A | .................... | .................... | .................... | .................... | 1 |
| B | 3171 hCoV-19/Scotland/CVR2988/2020\|EPI_ISL_439004\|2020- | .................... | ...................A | .................... | .................... | .................... | .................... | 1 |
| B | 3172 hCoV-19/Scotland/CVR2989/2020\|EPI_ISL_439005\|2020- | .................... | ...................A | .................... | .................... | .................... | .................... | 1 |
| B | 3173 hCoV-19/Scotland/CVR2990/2020\|EPI_ISL_439006\|2020- | .................... | .................... | .................... | .................... | .................... | .................... | |
| B | 3174 hCoV-19/Scotland/CVR3014/2020\|EPI_ISL_439007\|2020- | .................... | .................... | .................... | .................... | .................... | .................... | |
| B | 3175 hCoV-19/Scotland/CVR3019/2020\|EPI_ISL_439009\|2020- | .................... | .................... | .................... | .................... | .................... | .................... | |
| B | 3176 hCoV-19/Scotland/CVR3022/2020\|EPI_ISL_439012\|2020- | .................... | .................... | .................... | .................... | .................... | .................... | |
| B | 3177 hCoV-19/Scotland/CVR3023/2020\|EPI_ISL_439013\|2020- | .................... | .................... | .................... | .................... | .................... | .................... | |
| B | 3178 hCoV-19/Scotland/CVR3024/2020\|EPI_ISL_439014\|2020- | .................... | .................... | .................... | .................... | .................... | .................... | |
| B | 3179 hCoV-19/Scotland/CVR3025/2020\|EPI_ISL_439015\|2020- | .................... | .................... | .................... | .................... | .................... | .................... | |
| B | 3180 hCoV-19/Scotland/CVR3026/2020\|EPI_ISL_439016\|2020- | .................... | .................... | .................... | .................... | .................... | .................... | |
| B | 3181 hCoV-19/Scotland/CVR3028/2020\|EPI_ISL_439018\|2020- | .................... | .................... | .................... | .................... | .................... | .................... | |
| B | 3182 hCoV-19/Scotland/CVR3030/2020\|EPI_ISL_439020\|2020- | .................... | .................... | .................... | .................... | .................... | .................... | |
| B | 3183 hCoV-19/Scotland/CVR3035/2020\|EPI_ISL_439022\|2020- | .................... | .................... | .................... | .................... | .................... | .................... | |
| B | 3184 hCoV-19/Scotland/CVR306/2020\|EPI_ISL_439024\|2020-0 | .................... | ...................A | .................... | .................... | .................... | .................... | 1 |
| B | 3185 hCoV-19/Scotland/CVR3085/2020\|EPI_ISL_439026\|2020- | .................... | .................... | .................... | .................... | .................... | .................... | |
| B | 3186 hCoV-19/Scotland/CVR3086/2020\|EPI_ISL_439027\|2020- | .................... | .................... | .................... | .................... | .................... | .................... | |
| B | 3187 hCoV-19/Scotland/CVR3087/2020\|EPI_ISL_439028\|2020- | .................... | ...................A | .................... | .................... | .................... | .................... | 1 |
| B | 3188 hCoV-19/Scotland/CVR3088/2020\|EPI_ISL_439029\|2020- | .................... | .................... | .................... | .................... | .................... | .................... | |
| B | 3189 hCoV-19/England/BRIS-122043/2020\|EPI_ISL_440001\|20 | .................... | .................... | .................... | .................... | .................... | .................... | |
| B | 3190 hCoV-19/England/BRIS-12222F/2020\|EPI_ISL_440002\|20 | .................... | .................... | .................... | .................... | .................... | .................... | |
| B | 3191 hCoV-19/England/CAMB-73660/2020\|EPI_ISL_440004\|202 | .................... | .................... | .................... | .................... | .................... | .................... | |
| B | 3192 hCoV-19/England/BRIS-122122/2020\|EPI_ISL_440005\|20 | .................... | .................... | .................... | .................... | .................... | .................... | |
| B | 3193 hCoV-19/England/BRIS-121E2C/2020\|EPI_ISL_440006\|20 | .................... | .................... | .................... | .................... | .................... | .................... | |
| B | 3194 hCoV-19/England/CAMB-734EE/2020\|EPI_ISL_440007\|202 | .................... | .................... | .................... | .................... | .................... | .................... | |
| B | 3195 hCoV-19/England/BRIS-121DE3/2020\|EPI_ISL_440008\|20 | .................... | .................... | .................... | .................... | .................... | .................... | |
| B | 3196 hCoV-19/England/CAMB-72D7B/2020\|EPI_ISL_440009\|202 | .................... | .................... | .................... | .................... | .................... | .................... | |
| B | 3197 hCoV-19/England/CAMB-72C50/2020\|EPI_ISL_440010\|202 | .................... | .................... | .................... | .................... | .................... | .................... | |
| B | 3198 hCoV-19/England/CAMB-72DD5/2020\|EPI_ISL_440011\|202 | .................... | .................... | .................... | .................... | .................... | .................... | |
| B | 3199 hCoV-19/England/CAMB-7A0F2/2020\|EPI_ISL_441300\|202 | .................... | .................... | .................... | .................... | .................... | .................... | |
| B | 3200 hCoV-19/England/CAMB-79CB3/2020\|EPI_ISL_441301\|202 | .................... | .................... | .................... | .................... | .................... | .................... | |
| B | 3201 hCoV-19/England/CAMB-79925/2020\|EPI_ISL_441302\|202 | .................... | .................... | .................... | .................... | .................... | .................... | |
| B | 3202 hCoV-19/England/CAMB-7A65A/2020\|EPI_ISL_441303\|202 | .................... | .A.................. | .................... | .................... | .................... | .................... | |
| B | 3203 hCoV-19/England/CAMB-7A292/2020\|EPI_ISL_441304\|202 | .................... | .................... | .................... | .................... | .................... | .................... | |
| B | 3204 hCoV-19/England/CAMB-79B6B/2020\|EPI_ISL_441305\|202 | .................... | .................... | .................... | .................... | .................... | .................... | |

| | | | | | | | |
|---|---|---|---|---|---|---|---|
| B | 3205 hCoV-19/England/CAMB-79907/2020\|EPI_ISL_441307\|202 | ···················· | ···················· | ····················· | ···················· | ···················· | ···················· |
| B | 3206 hCoV-19/England/CAMB-79CD1/2020\|EPI_ISL_441308\|202 | ···················· | ···················· | ····················· | ···················· | ···················· | ···················· |
| B | 3207 hCoV-19/England/CAMB-7A03E/2020\|EPI_ISL_441309\|202 | ···················· | ···················· | ····················· | ···················· | ···················· | ···················· |
| B | 3208 hCoV-19/England/CAMB-79B7A/2020\|EPI_ISL_441310\|202 | ···················· | ···················· | ····················· | ···················· | ···················· | ···················· |
| B | 3209 hCoV-19/England/CAMB-79FBA/2020\|EPI_ISL_441311\|202 | ···················· | ···················· | ····················· | ···················· | ···················· | ···················· |
| B | 3210 hCoV-19/England/CAMB-77E46/2020\|EPI_ISL_441312\|202 | ···················· | ···················· | ····················· | ···················· | ···················· | ···················· |
| B | 3211 hCoV-19/England/CAMB-79D47/2020\|EPI_ISL_441313\|202 | ···················· | ···················· | ····················· | ···················· | ···················· | ···················· |
| B | 3212 hCoV-19/England/CAMB-7A274/2020\|EPI_ISL_441314\|202 | ···················· | ···················· | ····················· | ···················· | ···················· | ···················· |
| B | 3213 hCoV-19/England/CAMB-7A335/2020\|EPI_ISL_441315\|202 | ···················· | ···················· | ····················· | ···················· | ···················· | ···················· |
| B | 3214 hCoV-19/England/CAMB-7A669/2020\|EPI_ISL_441317\|202 | ···················· | ···················· | ····················· | ···················· | ···················· | ···················· |
| B | 3215 hCoV-19/England/CAMB-79F14/2020\|EPI_ISL_441319\|202 | ···················· | ···················· | ····················· | ···················· | ···················· | ···················· |
| B | 3216 hCoV-19/England/CAMB-7A371/2020\|EPI_ISL_441320\|202 | ···················· | ···················· | ····················· | ···················· | ···················· | ···················· |
| B | 3217 hCoV-19/England/CAMB-7A1D1/2020\|EPI_ISL_441321\|202 | ···················· | ···················· | ····················· | ···················· | ···················· | ···················· |
| B | 3218 hCoV-19/England/CAMB-7908A/2020\|EPI_ISL_441322\|202 | ···················· | ···················· | ····················· | ···················· | ···················· | ···················· |
| B | 3219 hCoV-19/England/CAMB-79EDB/2020\|EPI_ISL_441325\|202 | ···················· | ···················· | ····················· | ···················· | ···················· | ···················· |
| B | 3220 hCoV-19/England/CAMB-7A520/2020\|EPI_ISL_441326\|202 | ···················· | ···················· | ····················· | ···················· | ···················· | ···················· |
| B | 3221 hCoV-19/England/CAMB-79E71/2020\|EPI_ISL_441328\|202 | ···················· | ···················· | ····················· | ···················· | ···················· | ···················· |
| B | 3222 hCoV-19/England/CAMB-7A20B/2020\|EPI_ISL_441331\|202 | ···················· | ···················· | ····················· | ···················· | ···················· | ···················· |
| B | 3223 hCoV-19/England/CAMB-79D0B/2020\|EPI_ISL_441332\|202 | ···················· | ···················· | ····················· | ···················· | ···················· | ···················· |
| B | 3224 hCoV-19/England/CAMB-7A6B4/2020\|EPI_ISL_441333\|202 | ···················· | ·A·················· | ····················· | ···················· | ···················· | ···················· |
| B | 3225 hCoV-19/England/CAMB-7A405/2020\|EPI_ISL_441334\|202 | ···················· | ···················· | ····················· | ···················· | ···················· | ···················· |
| B | 3226 hCoV-19/England/CAMB-79B4D/2020\|EPI_ISL_441335\|202 | ···················· | ···················· | ····················· | ···················· | ···················· | ···················· |
| B | 3227 hCoV-19/England/CAMB-7A04D/2020\|EPI_ISL_441337\|202 | ···················· | ···················· | ····················· | ···················· | ···················· | ···················· |
| B | 3228 hCoV-19/England/CAMB-7A195/2020\|EPI_ISL_441338\|202 | ···················· | ···················· | ····················· | ···················· | ···················· | ···················· |
| B | 3229 hCoV-19/England/CAMB-77F70/2020\|EPI_ISL_441339\|202 | ···················· | ···················· | ····················· | ···················· | ···················· | ···················· |
| B | 3230 hCoV-19/England/CAMB-79F7E/2020\|EPI_ISL_441340\|202 | ···················· | ···················· | ····················· | ···················· | ···················· | ···················· |
| B | 3231 hCoV-19/England/CAMB-79E53/2020\|EPI_ISL_441341\|202 | ···················· | ···················· | ····················· | ···················· | ···················· | ···················· |
| B | 3232 hCoV-19/England/CAMB-7A9D9/2020\|EPI_ISL_441342\|202 | ···················· | ···················· | ····················· | ···················· | ···················· | ···················· |
| B | 3233 hCoV-19/England/CAMB-7EC18/2020\|EPI_ISL_442601\|202 | ···················· | ···················· | ····················· | ···················· | ···················· | ···················· |
| B | 3234 hCoV-19/England/CAMB-81C03/2020\|EPI_ISL_442602\|202 | ···················· | ···················· | ····················· | ···················· | ···················· | ···················· |
| B | 3235 hCoV-19/England/CAMB-81DB5/2020\|EPI_ISL_442603\|202 | ···················· | ···················· | ····················· | ···················· | ···················· | ···················· |
| B | 3236 hCoV-19/England/CAMB-81A63/2020\|EPI_ISL_442604\|202 | ···················· | ···················· | ····················· | ···················· | ···················· | ···················· |
| B | 3237 hCoV-19/England/CAMB-81B7F/2020\|EPI_ISL_442605\|202 | ···················· | ···················· | ····················· | ···················· | ···················· | ···················· |
| B | 3238 hCoV-19/England/CAMB-81720/2020\|EPI_ISL_442606\|202 | ···················· | ···················· | ····················· | ···················· | ···················· | ···················· |
| B | 3239 hCoV-19/England/CAMB-7EAA5/2020\|EPI_ISL_442607\|202 | ···················· | ···················· | ····················· | ···················· | ···················· | ···················· |
| B | 3240 hCoV-19/England/BRIS-12324C/2020\|EPI_ISL_442632\|20 | ···················· | ···················· | ····················· | ···················· | ···················· | ···················· |
| B | 3241 hCoV-19/England/BRIS-123ADB/2020\|EPI_ISL_442633\|20 | ···················· | ···················· | ····················· | ···················· | ···················· | ···················· |
| B | 3242 hCoV-19/England/BRIS-1238A4/2020\|EPI_ISL_442634\|20 | ···················· | ····A··············· | ····················· | ···················· | ···················· | ···················· |
| B | 3243 hCoV-19/England/BRIS-12316D/2020\|EPI_ISL_442636\|20 | ···················· | ···················· | ····················· | ···················· | ···················· | ···················· |
| B | 3244 hCoV-19/England/BRIS-123B14/2020\|EPI_ISL_442637\|20 | ···················· | ····A··············· | ····················· | ···················· | ···················· | ···················· |
| B | 3245 hCoV-19/England/BRIS-12381D/2020\|EPI_ISL_442638\|20 | ···················· | ···················· | ····················· | ···················· | ···················· | ···················· |
| B | 3246 hCoV-19/England/BRIS-123B7E/2020\|EPI_ISL_442639\|20 | ···················· | ···················· | ····················· | ···················· | ···················· | ···················· |
| B | 3247 hCoV-19/England/BRIS-1236B9/2020\|EPI_ISL_442640\|20 | ···················· | ···················· | ····················· | ···················· | ···················· | ···················· |
| B | 3248 hCoV-19/England/BRIS-1235BC/2020\|EPI_ISL_442641\|20 | ···················· | ···················· | ····················· | ···················· | ···················· | ···················· |
| B | 3249 hCoV-19/England/BRIS-123288/2020\|EPI_ISL_442643\|20 | ···················· | ···················· | ····················· | ···················· | ···················· | ···················· |
| B | 3250 hCoV-19/England/BRIS-12391A/2020\|EPI_ISL_442644\|20 | ········T··········· | ···················· | ····················· | ···················· | ···················· | ···················· |
| B | 3251 hCoV-19/England/BRIS-123437/2020\|EPI_ISL_442645\|20 | ···················· | ···················· | ····················· | ···················· | ···················· | ···················· |
| B | 3252 hCoV-19/England/BRIS-123631/2020\|EPI_ISL_442646\|20 | ···················· | ···················· | ····················· | ···················· | ···················· | ···················· |
| B | 3253 hCoV-19/England/BRIS-123BC9/2020\|EPI_ISL_442647\|20 | ···················· | ···················· | ····················· | ···················· | ···················· | ···················· |
| B | 3254 hCoV-19/England/BRIS-123C20/2020\|EPI_ISL_442648\|20 | ···················· | ···················· | ····················· | ···················· | ···················· | ···················· |
| B | 3255 hCoV-19/England/BRIS-123AF9/2020\|EPI_ISL_442649\|20 | ···················· | ·A·················· | ····················· | ···················· | ···················· | ···················· |
| B | 3256 hCoV-19/England/BRIS-123710/2020\|EPI_ISL_442650\|20 | ···················· | ···················· | ····················· | ···················· | ···················· | ···················· |
| B | 3257 hCoV-19/England/BRIS-123886/2020\|EPI_ISL_442652\|20 | ···················· | ···················· | ····················· | ···················· | ···················· | ···················· |
| B | 3258 hCoV-19/England/BRIS-123570/2020\|EPI_ISL_442653\|20 | ···················· | ···················· | ····················· | ···················· | ···················· | ···················· |
| B | 3259 hCoV-19/England/BRIS-1239DE/2020\|EPI_ISL_442654\|20 | ···················· | ···················· | ····················· | ···················· | ···················· | ···················· |
| B | 3260 hCoV-19/England/BRIS-1235DA/2020\|EPI_ISL_442655\|20 | ···················· | ···················· | ····················· | ···················· | ···················· | ···················· |
| B | 3261 hCoV-19/England/BRIS-123965/2020\|EPI_ISL_442656\|20 | ···················· | ···················· | ····················· | ···················· | ···················· | ···················· |
| B | 3262 hCoV-19/England/BRIS-123464/2020\|EPI_ISL_442657\|20 | ···················· | ···················· | ····················· | ···················· | ···················· | ···················· |
| B | 3263 hCoV-19/England/BRIS-1232C4/2020\|EPI_ISL_442658\|20 | ···················· | ···················· | ····················· | ···················· | ···················· | ···················· |
| B | 3264 hCoV-19/England/BRIS-1236D7/2020\|EPI_ISL_442660\|20 | ···················· | ···················· | ····················· | ···················· | ···················· | ···················· |
| B | 3265 hCoV-19/England/BRIS-1230E8/2020\|EPI_ISL_442662\|20 | ···················· | ···················· | ····················· | ···················· | ···················· | ···················· |
| B | 3266 hCoV-19/England/BRIS-12372F/2020\|EPI_ISL_442663\|20 | ···················· | ···················· | ····················· | ···················· | ···················· | ···················· |
| B | 3267 hCoV-19/England/BRIS-12365F/2020\|EPI_ISL_442664\|20 | ···················· | ···················· | ····················· | ···················· | ···················· | ···················· |
| B | 3268 hCoV-19/England/BRIS-123349/2020\|EPI_ISL_442665\|20 | ···················· | ···················· | ····················· | ···················· | ···················· | ···················· |
| B | 3269 hCoV-19/England/BRIS-123C4E/2020\|EPI_ISL_442667\|20 | ···················· | ···················· | ····················· | ···················· | ···················· | ···················· |
| B | 3270 hCoV-19/England/BRIS-123428/2020\|EPI_ISL_442668\|20 | ···················· | ···················· | ····················· | ···················· | ···················· | ···················· |
| B | 3271 hCoV-19/England/BRIS-123938/2020\|EPI_ISL_442669\|20 | ···················· | ···················· | ····················· | ···················· | ···················· | ···················· |
| B | 3272 hCoV-19/England/BRIS-123929/2020\|EPI_ISL_442670\|20 | ········T··········· | ···················· | ····················· | ···················· | ···················· | ···················· |
| B | 3273 hCoV-19/England/BRIS-12317C/2020\|EPI_ISL_442671\|20 | ···················· | ···················· | ····················· | ···················· | ···················· | ···················· |
| B | 3274 hCoV-19/England/BRIS-123C8A/2020\|EPI_ISL_442672\|20 | ···················· | ···················· | ····················· | ···················· | ···················· | ···················· |
| B | 3275 hCoV-19/England/BRIS-1239B0/2020\|EPI_ISL_442673\|20 | ···················· | ···················· | ····················· | ···················· | ···················· | ···················· |
| B | 3276 hCoV-19/England/BRIS-1236AA/2020\|EPI_ISL_442674\|20 | ···················· | ···················· | ····················· | ···················· | ···················· | ···················· |
| B | 3277 hCoV-19/England/BRIS-125CC4/2020\|EPI_ISL_443922\|20 | ···················· | ···················· | ····················· | ···················· | ···················· | ···················· |
| B | 3278 hCoV-19/England/BRIS-125A51/2020\|EPI_ISL_443924\|20 | ···················· | ···················· | ····················· | ···················· | ···················· | ···················· |
| B | 3279 hCoV-19/England/BRIS-124CB6/2020\|EPI_ISL_443925\|20 | ···················· | ···················· | ····················· | ···················· | ···················· | ···················· |
| B | 3280 hCoV-19/England/BRIS-12557E/2020\|EPI_ISL_443927\|20 | ···················· | ···················· | ····················· | ···················· | ···················· | ···················· |
| B | 3281 hCoV-19/England/BRIS-125CA6/2020\|EPI_ISL_443933\|20 | ···················· | ···················· | ····················· | ···················· | ···················· | ···················· |
| B | 3282 hCoV-19/England/BRIS-125A8E/2020\|EPI_ISL_443935\|20 | ···················· | ···················· | ····················· | ···················· | ···················· | ···················· |

| | | | | | | | | |
|---|---|---|---|---|---|---|---|---|
| B | 3283 | hCoV-19/England/BRIS-12549F/2020\|EPI_ISL_443936\|20 | .................... | .................... | .................... | .................... | .................... | .................... |
| B | 3284 | hCoV-19/England/BRIS-1251B6/2020\|EPI_ISL_443938\|20 | .................... | .................... | .................... | .................... | .................... | .................... |
| B | 3285 | hCoV-19/England/BRIS-124CF2/2020\|EPI_ISL_443939\|20 | .................... | .................... | .................... | .................... | .................... | .................... |
| B | 3286 | hCoV-19/England/BRIS-125F43/2020\|EPI_ISL_443962\|20 | .................... | .................... | .................... | .................... | .................... | .................... |
| B | 3287 | hCoV-19/England/BRIS-12580C/2020\|EPI_ISL_443963\|20 | .................... | .................... | .................... | .................... | .................... | .................... |
| B | 3288 | hCoV-19/England/BRIS-124B04/2020\|EPI_ISL_443964\|20 | .................... | .................... | .................... | .................... | .................... | .................... |
| B | 3289 | hCoV-19/England/BRIS-125C5B/2020\|EPI_ISL_443967\|20 | .................... | .................... | .................... | .................... | .................... | .................... |
| B | 3290 | hCoV-19/England/BRIS-125972/2020\|EPI_ISL_443968\|20 | .................... | .................... | .................... | .................... | .................... | .................... |
| B | 3291 | hCoV-19/England/BRIS-12531A/2020\|EPI_ISL_443969\|20 | .................... | .................... | .................... | .................... | .................... | .................... |
| B | 3292 | hCoV-19/England/BRIS-1257C3/2020\|EPI_ISL_443970\|20 | .................... | .................... | .................... | .................... | .................... | .................... |
| B | 3293 | hCoV-19/England/BRIS-125444/2020\|EPI_ISL_443971\|20 | .................... | .................... | .................... | .................... | .................... | .................... |
| B | 3294 | hCoV-19/England/BRIS-12556F/2020\|EPI_ISL_443972\|20 | .................... | .................... | .................... | .................... | .................... | .................... |
| B | 3295 | hCoV-19/England/BRIS-1257E1/2020\|EPI_ISL_443973\|20 | .................... | .................... | .................... | .................... | .................... | .................... |
| B | 3296 | hCoV-19/England/BRIS-1249CE/2020\|EPI_ISL_443975\|20 | .................... | .................... | .................... | .................... | .................... | .................... |
| B | 3297 | hCoV-19/England/BRIS-124EA1/2020\|EPI_ISL_443976\|20 | .................... | .................... | .................... | .................... | .................... | .................... |
| B | 3298 | hCoV-19/England/BRIS-125A06/2020\|EPI_ISL_443978\|20 | .................... | .................... | .................... | .................... | .................... | .................... |
| B | 3299 | hCoV-19/England/BRIS-125611/2020\|EPI_ISL_443980\|20 | .................... | .................... | .................... | .................... | .................... | .................... |
| B | 3300 | hCoV-19/England/BRIS-125426/2020\|EPI_ISL_443981\|20 | .................... | .................... | .................... | .................... | .................... | .................... |
| B | 3301 | hCoV-19/England/BRIS-124E83/2020\|EPI_ISL_443982\|20 | .................... | .................... | .................... | .................... | .................... | .................... |
| B | 3302 | hCoV-19/England/BRIS-12582A/2020\|EPI_ISL_443983\|20 | .................... | .................... | .................... | .................... | .................... | .................... |
| B | 3303 | hCoV-19/England/BRIS-124ABC/2020\|EPI_ISL_443985\|20 | .................... | .................... | .................... | .................... | .................... | .................... |
| B | 3304 | hCoV-19/England/BRIS-1247B5/2020\|EPI_ISL_443986\|20 | .................... | .................... | .................... | .................... | .................... | .................... |
| B | 3305 | hCoV-19/England/BRIS-125BE5/2020\|EPI_ISL_443987\|20 | .................... | .................... | .................... | .................... | .................... | .................... |
| B | 3306 | hCoV-19/England/BRIS-124BC8/2020\|EPI_ISL_443988\|20 | .................... | .................... | .................... | .................... | .................... | .................... |
| B | 3307 | hCoV-19/England/BRIS-12565D/2020\|EPI_ISL_443989\|20 | .................... | .................... | .................... | .................... | .................... | .................... |
| B | 3308 | hCoV-19/England/BRIS-124E74/2020\|EPI_ISL_443990\|20 | .................... | .................... | .................... | .................... | .................... | .................... |
| B | 3309 | hCoV-19/England/BRIS-12506E/2020\|EPI_ISL_443991\|20 | .................... | .................... | .................... | .................... | .................... | .................... |
| B | 3310 | hCoV-19/England/BRIS-125D94/2020\|EPI_ISL_443992\|20 | .................... | .................... | .................... | .................... | .................... | .................... |
| B | 3311 | hCoV-19/England/BRIS-1253ED/2020\|EPI_ISL_443996\|20 | .................... | .................... | .................... | .................... | .................... | .................... |
| B | 3312 | hCoV-19/England/BRIS-124E1A/2020\|EPI_ISL_443997\|20 | .................... | .................... | .................... | .................... | .................... | .................... |
| B | 3313 | hCoV-19/England/BRIS-1250B9/2020\|EPI_ISL_443998\|20 | .................... | .................... | .................... | .................... | .................... | .................... |
| B | 3314 | hCoV-19/England/BRIS-124F53/2020\|EPI_ISL_443999\|20 | .................... | .................... | .................... | .................... | .................... | .................... |
| B | 3315 | hCoV-19/Wales/PHWC-2EB92/2020\|EPI_ISL_446121\|2020- | .................... | .A.................. | .................... | .................... | .................... | .................... |
| B | 3316 | hCoV-19/Wales/PHWC-2EBDE/2020\|EPI_ISL_446124\|2020- | .................... | ....A............... | .................... | .................... | .................... | .................... |
| B | 3317 | hCoV-19/Wales/PHWC-2EBED/2020\|EPI_ISL_446125\|2020- | .................... | .................... | .................... | .................... | .................... | .................... |
| B | 3318 | hCoV-19/Wales/PHWC-2EBFC/2020\|EPI_ISL_446126\|2020- | .................... | .................... | .................... | .................... | .................... | .................... |
| B | 3319 | hCoV-19/Wales/PHWC-2EC35/2020\|EPI_ISL_446130\|2020- | .................... | .................... | .................... | .................... | .................... | .................... |
| B | 3320 | hCoV-19/Wales/PHWC-2EC44/2020\|EPI_ISL_446131\|2020- | .................... | .................... | .................... | .................... | .................... | .................... |
| B | 3321 | hCoV-19/Wales/PHWC-2ECDB/2020\|EPI_ISL_446139\|2020- | .................... | ....A............... | .................... | .................... | .................... | .................... |
| B | 3322 | hCoV-19/Wales/PHWC-2ED14/2020\|EPI_ISL_446143\|2020- | .................... | .................... | .................... | .................... | .................... | .................... |
| B | 3323 | hCoV-19/Wales/PHWC-2ED23/2020\|EPI_ISL_446144\|2020- | .................... | .................... | .................... | .................... | .................... | .................... |
| B | 3324 | hCoV-19/Wales/PHWC-2ED41/2020\|EPI_ISL_446146\|2020- | .................... | .............A...... | .................... | .................... | .................... | .................... |
| B | 3325 | hCoV-19/Wales/PHWC-2ED50/2020\|EPI_ISL_446147\|2020- | .................... | .................... | .................... | .................... | .................... | .................... |
| B | 3326 | hCoV-19/Wales/PHWC-2ED9C/2020\|EPI_ISL_446151\|2020- | .................... | .................... | .................... | .................... | .................... | .................... |
| B | 3327 | hCoV-19/Wales/PHWC-2EDBA/2020\|EPI_ISL_446153\|2020- | .................... | .................... | .................... | .................... | .................... | .................... |
| B | 3328 | hCoV-19/Wales/PHWC-2EDE7/2020\|EPI_ISL_446156\|2020- | .................... | .................... | .................... | .................... | .................... | .................... |
| B | 3329 | hCoV-19/USA/NY-NYUMC308/2020\|EPI_ISL_430335\|2020-0 | .................... | .................... | .................... | .................... | .................... | .................... |
| B | 3330 | hCoV-19/USA/NY-NYUMC309/2020\|EPI_ISL_430336\|2020-0 | .................... | .................... | .................... | .................... | .................... | .................... |
| B | 3331 | hCoV-19/USA/NY-NYUMC328/2020\|EPI_ISL_430355\|2020-0 | .................... | .................... | .................... | .................... | .................... | .................... |
| B | 3332 | hCoV-19/USA/NJ-NYUMC329/2020\|EPI_ISL_430356\|2020-0 | .................... | .................... | .................... | .................... | .................... | .................... |
| B | 3333 | hCoV-19/USA/NY-NYUMC330/2020\|EPI_ISL_430357\|2020-0 | .................... | .................... | .................... | .................... | .................... | .................... |
| B | 3334 | hCoV-19/USA/NY-NYUMC331/2020\|EPI_ISL_430358\|2020-0 | .................... | .................... | .................... | .................... | .................... | .................... |
| B | 3335 | hCoV-19/USA/NY-NYUMC332/2020\|EPI_ISL_430359\|2020-0 | .................... | .................... | .................... | .................... | .................... | .................... |
| B | 3336 | hCoV-19/USA/NY-NYUMC333/2020\|EPI_ISL_430360\|2020-0 | .................... | .................... | .................... | .................... | .................... | .................... |
| B | 3337 | hCoV-19/USA/NY-NYUMC334/2020\|EPI_ISL_430361\|2020-0 | .................... | .................... | .................... | .................... | .................... | .................... |
| B | 3338 | hCoV-19/USA/NY-NYUMC335/2020\|EPI_ISL_430362\|2020-0 | .................... | .................... | .................... | .................... | .................... | .................... |
| B | 3339 | hCoV-19/USA/NY-NYUMC336/2020\|EPI_ISL_430363\|2020-0 | .................... | .................... | .................... | .................... | .................... | .................... |
| B | 3340 | hCoV-19/USA/NY-NYUMC337/2020\|EPI_ISL_430364\|2020-0 | .................... | .................... | .................... | .................... | .................... | .................... |
| B | 3341 | hCoV-19/USA/NY-NYUMC338/2020\|EPI_ISL_430365\|2020-0 | .................... | .................... | .................... | .................... | .................... | .................... |
| B | 3342 | hCoV-19/USA/NY-NYUMC339/2020\|EPI_ISL_430366\|2020-0 | .................... | .................... | .................... | .................... | .................... | .................... |
| B | 3343 | hCoV-19/USA/NY-NYUMC340/2020\|EPI_ISL_430367\|2020-0 | .................... | .................... | .................... | .................... | .................... | .................... |
| B | 3344 | hCoV-19/USA/NY-NYUMC341/2020\|EPI_ISL_430368\|2020-0 | .................... | .................... | .................... | .................... | .................... | .................... |
| B | 3345 | hCoV-19/USA/NY-NYUMC342/2020\|EPI_ISL_430369\|2020-0 | .................... | .................... | .................... | .................... | .................... | .................... |
| B | 3346 | hCoV-19/USA/NY-NYUMC343/2020\|EPI_ISL_430370\|2020-0 | .................... | .................... | .................... | .................... | .................... | .................... |
| B | 3347 | hCoV-19/USA/NJ-NYUMC344/2020\|EPI_ISL_430371\|2020-0 | .................... | .................... | .................... | .................... | .................... | .................... |
| B | 3348 | hCoV-19/USA/NY-NYUMC345/2020\|EPI_ISL_430372\|2020-0 | .................... | .................... | .................... | .................... | .................... | .................... |
| B | 3349 | hCoV-19/USA/NY-NYUMC346/2020\|EPI_ISL_430373\|2020-0 | .................... | .................... | .................... | .................... | .................... | .................... |
| B | 3350 | hCoV-19/USA/NY-NYUMC347/2020\|EPI_ISL_430374\|2020-0 | .................... | .................... | .................... | .................... | .................... | .................... |
| B | 3351 | hCoV-19/USA/NY-NYUMC348/2020\|EPI_ISL_430375\|2020-0 | .................... | .................... | .................... | .................... | .................... | .................... |
| B | 3352 | hCoV-19/USA/NY-NYUMC349/2020\|EPI_ISL_430376\|2020-0 | .................... | .................... | .................... | .................... | .................... | .................... |
| B | 3354 | hCoV-19/Australia/VIC1135/2020\|EPI_ISL_430601\|2020 | .................... | .................... | .................... | .................... | .................... | .................... |
| B | 3356 | hCoV-19/Australia/VIC1159/2020\|EPI_ISL_430604\|2020 | .................... | .................... | .................... | .................... | .................... | .................... |
| B | 3360 | hCoV-19/USA/WA-S340/2020\|EPI_ISL_434100\|2020-04-04 | .................... | .................... | .................... | .................... | .................... | .................... |
| B | 3361 | hCoV-19/USA/WA-S341/2020\|EPI_ISL_434101\|2020-04-02 | .................... | .................... | .................... | .................... | .................... | .................... |
| B | 3362 | hCoV-19/USA/WA-S342/2020\|EPI_ISL_434102\|2020-04-02 | .................... | .................... | .................... | .................... | .................... | .................... |
| B | 3363 | hCoV-19/USA/WA-S343/2020\|EPI_ISL_434103\|2020-04-02 | .................... | .................... | .................... | .................... | .................... | .................... |
| B | 3364 | hCoV-19/USA/WA-S344/2020\|EPI_ISL_434104\|2020-04-02 | .................... | .................... | .................... | .................... | .................... | .................... |
| B | 3365 | hCoV-19/USA/WA-S345/2020\|EPI_ISL_434105\|2020-04-05 | .................... | .................... | .................... | .................... | .................... | .................... |

| | | | | | | | | |
|---|---|---|---|---|---|---|---|---|
| B | 3366 | hCoV-19/USA/WA-S346/2020\|EPI_ISL_434106\|2020-04-04 | ···················· | ···················· | ···················· | ···················· | ···················· | ···················· |
| B | 3367 | hCoV-19/USA/WA-S347/2020\|EPI_ISL_434107\|2020-04-01 | ···················· | ···················· | ···················· | ···················· | ···················· | ···················· |
| B | 3368 | hCoV-19/USA/WA-S348/2020\|EPI_ISL_434108\|2020-04-01 | ···················· | ···················· | ···················· | ···················· | ···················· | ···················· |
| B | 3369 | hCoV-19/USA/WA-S349/2020\|EPI_ISL_434109\|2020-04-02 | ···················· | ···················· | ···················· | ···················· | ···················· | ···················· |
| B | 3370 | hCoV-19/USA/WA-S350/2020\|EPI_ISL_434110\|2020-04-02 | ···················· | ···················· | ···················· | ···················· | ···················· | ···················· |
| B | 3371 | hCoV-19/USA/WA-S351/2020\|EPI_ISL_434111\|2020-03-25 | ···················· | ···················· | ···················· | ···················· | ···················· | ···················· |
| B | 3372 | hCoV-19/USA/WA-S352/2020\|EPI_ISL_434112\|2020-03-25 | ···················· | ···················· | ···················· | ···················· | ···················· | ···················· |
| B | 3373 | hCoV-19/USA/WA-S353/2020\|EPI_ISL_434113\|2020-03-25 | ···················· | ···················· | ···················· | ···················· | ···················· | ···················· |
| B | 3374 | hCoV-19/USA/WA-S354/2020\|EPI_ISL_434114\|2020-03-25 | ···················· | ···················· | ···················· | ···················· | ···················· | ···················· |
| B | 3375 | hCoV-19/USA/WA-S355/2020\|EPI_ISL_434115\|2020-03-25 | ···················· | ···················· | ···················· | ···················· | ···················· | ···················· |
| B | 3376 | hCoV-19/USA/WA-S356/2020\|EPI_ISL_434116\|2020-03-25 | ···················· | ···················· | ···················· | ···················· | ···················· | ···················· |
| B | 3377 | hCoV-19/USA/WA-S357/2020\|EPI_ISL_434117\|2020-03-25 | ···················· | ···················· | ···················· | ···················· | ···················· | ···················· |
| B | 3378 | hCoV-19/USA/WA-S358/2020\|EPI_ISL_434118\|2020-03-25 | ···················· | ···················· | ···················· | ···················· | ···················· | ···················· |
| B | 3379 | hCoV-19/USA/WA-S359/2020\|EPI_ISL_434119\|2020-03-25 | ···················· | ···················· | ···················· | ···················· | ···················· | ···················· |
| B | 3380 | hCoV-19/USA/WA-S360/2020\|EPI_ISL_434120\|2020-03-25 | ···················· | ···················· | ···················· | ···················· | ···················· | ···················· |
| B | 3381 | hCoV-19/USA/WA-S361/2020\|EPI_ISL_434121\|2020-03-26 | ···················· | ···················· | ···················· | ···················· | ···················· | ···················· |
| B | 3382 | hCoV-19/USA/WA-S362/2020\|EPI_ISL_434122\|2020-03-26 | ···················· | ···················· | ···················· | ···················· | ···················· | ···················· |
| B | 3383 | hCoV-19/USA/WA-S363/2020\|EPI_ISL_434123\|2020-03-26 | ···················· | ···················· | ···················· | ···················· | ···················· | ···················· |
| B | 3384 | hCoV-19/USA/WA-S364/2020\|EPI_ISL_434124\|2020-03-26 | ···················· | ···················· | ···················· | ···················· | ···················· | ···················· |
| B | 3385 | hCoV-19/USA/WA-S368/2020\|EPI_ISL_434128\|2020-03-26 | ···················· | ···················· | ···················· | ···················· | ···················· | ···················· |
| B | 3386 | hCoV-19/USA/WA-S369/2020\|EPI_ISL_434129\|2020-03-26 | ···················· | ···················· | ···················· | ···················· | ···················· | ···················· |
| B | 3387 | hCoV-19/USA/WA-S370/2020\|EPI_ISL_434130\|2020-03-28 | ···················· | ···················· | ···················· | ···················· | ···················· | ···················· |
| B | 3388 | hCoV-19/USA/WA-S371/2020\|EPI_ISL_434131\|2020-03-27 | ···················· | ···················· | ···················· | ···················· | ···················· | ···················· |
| B | 3389 | hCoV-19/USA/WA-S372/2020\|EPI_ISL_434132\|2020-03-26 | ···················· | ···················· | ···················· | ···················· | ···················· | ···················· |
| B | 3390 | hCoV-19/USA/WA-S373/2020\|EPI_ISL_434133\|2020-03-28 | ···················· | ···················· | ···················· | ···················· | ···················· | ···················· |
| B | 3391 | hCoV-19/USA/WA-S374/2020\|EPI_ISL_434134\|2020-03-26 | ···················· | ···················· | ···················· | ···················· | ···················· | ···················· |
| B | 3392 | hCoV-19/USA/WA-S375/2020\|EPI_ISL_434135\|2020-03-26 | ···················· | ···················· | ···················· | ···················· | ···················· | ···················· |
| B | 3393 | hCoV-19/USA/WA-S376/2020\|EPI_ISL_434136\|2020-03-27 | ···················· | ···················· | ···················· | ···················· | ···················· | ···················· |
| B | 3394 | hCoV-19/USA/WA-S377/2020\|EPI_ISL_434137\|2020-03-26 | ···················· | ···················· | ···················· | ···················· | ···················· | ···················· |
| B | 3395 | hCoV-19/USA/WA-S378/2020\|EPI_ISL_434138\|2020-03-27 | ···················· | ···················· | ···················· | ···················· | ···················· | ···················· |
| B | 3396 | hCoV-19/USA/WA-S391/2020\|EPI_ISL_434151\|2020-03-26 | ···················· | ···················· | ···················· | ···················· | ···················· | ···················· |
| B | 3397 | hCoV-19/USA/WA-S420/2020\|EPI_ISL_434180\|2020-03-28 | ···················· | ···················· | ···················· | ···················· | ···················· | ···················· |
| B | 3398 | hCoV-19/USA/WA-S422/2020\|EPI_ISL_434182\|2020-03-25 | ···················· | ···················· | ···················· | ···················· | ···················· | ···················· |
| B | 3399 | hCoV-19/USA/WA-S423/2020\|EPI_ISL_434183\|2020-03-28 | ···················· | ···················· | ···················· | ···················· | ···················· | ···················· |
| B | 3400 | hCoV-19/USA/WA-S424/2020\|EPI_ISL_434184\|2020-03-28 | ···················· | ···················· | ···················· | ···················· | ···················· | ···················· |
| B | 3401 | hCoV-19/USA/WA-S425/2020\|EPI_ISL_434185\|2020-03-28 | ···················· | ···················· | ···················· | ···················· | ···················· | ···················· |
| B | 3402 | hCoV-19/USA/WA-S426/2020\|EPI_ISL_434186\|2020-03-28 | ···················· | ···················· | ···················· | ···················· | ···················· | ···················· |
| B | 3403 | hCoV-19/USA/WA-S427/2020\|EPI_ISL_434187\|2020-03-28 | ···················· | ···················· | ···················· | ···················· | ···················· | ···················· |
| B | 3404 | hCoV-19/USA/WA-S428/2020\|EPI_ISL_434188\|2020-03-28 | ···················· | ···················· | ···················· | ···················· | ···················· | ···················· |
| B | 3405 | hCoV-19/USA/WA-S429/2020\|EPI_ISL_434189\|2020-03-25 | ···················· | ···················· | ···················· | ···················· | ···················· | ···················· |
| B | 3406 | hCoV-19/USA/WA-S430/2020\|EPI_ISL_434190\|2020-03-25 | ···················· | ···················· | ···················· | ···················· | ···················· | ···················· |
| B | 3407 | hCoV-19/USA/WA-S431/2020\|EPI_ISL_434191\|2020-03-28 | ···················· | ···················· | ···················· | ···················· | ···················· | ···················· |
| B | 3408 | hCoV-19/USA/WA-S432/2020\|EPI_ISL_434192\|2020-03-28 | ···················· | ···················· | ···················· | ···················· | ···················· | ···················· |
| B | 3409 | hCoV-19/USA/WA-S433/2020\|EPI_ISL_434193\|2020-03-28 | ···················· | ···················· | ···················· | ···················· | ···················· | ···················· |
| B | 3410 | hCoV-19/USA/WA-S434/2020\|EPI_ISL_434194\|2020-03-28 | ···················· | ···················· | ···················· | ···················· | ···················· | ···················· |
| B | 3411 | hCoV-19/USA/WA-S436/2020\|EPI_ISL_434196\|2020-03-31 | ···················· | ···················· | ···················· | ···················· | ···················· | ···················· |
| B | 3412 | hCoV-19/USA/WA-S437/2020\|EPI_ISL_434197\|2020-03-31 | ···················· | ···················· | ···················· | ···················· | ···················· | ···················· |
| B | 3413 | hCoV-19/USA/WA-S439/2020\|EPI_ISL_434199\|2020-04-02 | ···················· | ···················· | ···················· | ···················· | ················G··· | ···················· |
| B | 3414 | hCoV-19/USA/WI-GMF-01602/2020\|EPI_ISL_435400\|2020- | ···················· | ···················· | ···················· | ···················· | ···················· | ···················· |
| B | 3415 | hCoV-19/USA/WI-GMF-01828/2020\|EPI_ISL_435401\|2020- | ···················· | ···················· | ···················· | ···················· | ···················· | ···················· |
| B | 3416 | hCoV-19/USA/WI-GMF-01854/2020\|EPI_ISL_435402\|2020- | ···················· | ···················· | ···················· | ···················· | ···················· | ···················· |
| B | 3417 | hCoV-19/Hungary/SRC-00066/2020\|EPI_ISL_435403\|2020 | ···················· | ···················· | ···················· | ···················· | ···················· | ···················· |
| B | 3418 | hCoV-19/Hungary/SRC-00067/2020\|EPI_ISL_435404\|2020 | ···················· | ···················· | ···················· | ···················· | ···················· | ···················· |
| B | 3419 | hCoV-19/Hungary/SRC-00126/2020\|EPI_ISL_435405\|2020 | ···················· | ···················· | ···················· | ···················· | ···················· | ···················· |
| B | 3420 | hCoV-19/Hungary/SRC-00175/2020\|EPI_ISL_435406\|2020 | ···················· | ···················· | ···················· | ···················· | ···················· | ···················· |
| B | 3421 | hCoV-19/Hungary/SRC-00183/2020\|EPI_ISL_435407\|2020 | ···················· | ···················· | ···················· | ···················· | ···················· | ···················· |
| B | 3422 | hCoV-19/Hungary/SRC-00186/2020\|EPI_ISL_435408\|2020 | ···················· | ···················· | ···················· | ···················· | ···················· | ···················· |
| B | 3423 | hCoV-19/Hungary/SRC-00417/2020\|EPI_ISL_435409\|2020 | ···················· | ···················· | ···················· | ···················· | ···················· | ···················· |
| B | 3424 | hCoV-19/Hungary/SRC-00419/2020\|EPI_ISL_435410\|2020 | ···················· | ···················· | ···················· | ···················· | ···················· | ···················· |
| B | 3425 | hCoV-19/Hungary/SRC-00541/2020\|EPI_ISL_435411\|2020 | ···················· | ···················· | ···················· | ···················· | ···················· | ···················· |
| B | 3426 | hCoV-19/Hungary/SRC-00792/2020\|EPI_ISL_435412\|2020 | ···················· | ···················· | ···················· | ···················· | ···················· | ···················· |
| B | 3427 | hCoV-19/Hungary/SRC-00817/2020\|EPI_ISL_435413\|2020 | ···················· | ···················· | ···················· | ···················· | ···················· | ···················· |
| B | 3428 | hCoV-19/Hungary/SRC-00827/2020\|EPI_ISL_435414\|2020 | ···················· | ···················· | ···················· | ···················· | ···················· | ···················· |
| B | 3429 | hCoV-19/Hungary/SRC-00836/2020\|EPI_ISL_435415\|2020 | ···················· | ···················· | ···················· | ···················· | ···················· | ···················· |
| B | 3430 | hCoV-19/Hungary/SRC-00620/2020\|EPI_ISL_435416\|2020 | ···················· | ···················· | ···················· | ···················· | ···················· | ···················· |
| B | 3431 | hCoV-19/Hungary/SRC-00777/2020\|EPI_ISL_435417\|2020 | ···················· | ···················· | ···················· | ···················· | ···················· | ···················· |
| B | 3432 | hCoV-19/Hungary/SRC-01136/2020\|EPI_ISL_435418\|2020 | ···················· | ···················· | ···················· | ···················· | ···················· | ···················· |
| B | 3433 | hCoV-19/Hungary/SRC-00055/2020\|EPI_ISL_435419\|2020 | ···················· | ···················· | ···················· | ···················· | ···················· | ···················· |
| B | 3434 | hCoV-19/Hungary/MBL-002/2020\|EPI_ISL_435420\|2020-0 | ···················· | ···················· | ···················· | ···················· | ···················· | ···················· |
| B | 3435 | hCoV-19/Hungary/MBL-003/2020\|EPI_ISL_435421\|2020-0 | ···················· | ···················· | ···················· | ···················· | ···················· | ···················· |
| B | 3436 | hCoV-19/Hungary/MBL-464/2020\|EPI_ISL_435422\|2020-0 | ···················· | ···················· | ···················· | ···················· | ···················· | ···················· |
| B | 3437 | hCoV-19/Hungary/MBL-465/2020\|EPI_ISL_435423\|2020-0 | ···················· | ···················· | ···················· | ···················· | ···················· | ···················· |
| B | 3438 | hCoV-19/Hungary/MBL-469/2020\|EPI_ISL_435424\|2020-0 | ···················· | ···················· | ···················· | ···················· | ···················· | ···················· |
| B | 3439 | hCoV-19/Hungary/SRC-17881c/2020\|EPI_ISL_435425\|202 | ···················· | ···················· | ···················· | ···················· | ···················· | ···················· |
| B | 3440 | hCoV-19/Hungary/SRC-00105w/2020\|EPI_ISL_435426\|202 | ···················· | ···················· | ···················· | ···················· | ···················· | ···················· |
| B | 3441 | hCoV-19/Hungary/SRC-00278w/2020\|EPI_ISL_435427\|202 | ···················· | ···················· | ···················· | ···················· | ···················· | ···················· |
| B | 3442 | hCoV-19/Hungary/SRC-02801w/2020\|EPI_ISL_435428\|202 | ···················· | ···················· | ···················· | ···················· | ···················· | ···················· |
| B | 3443 | hCoV-19/Hungary/SRC-03597w/2020\|EPI_ISL_435429\|202 | ···················· | ···················· | ···················· | ···················· | ···················· | ···················· |

| | | | | | | | |
|---|---|---|---|---|---|---|---|
| B | 3444 hCoV-19/Hungary/SRC-03670w/2020\|EPI_ISL_435430\|202 | ···················· | ···················· | ····················· | ···················· | ···················· | ···················· |
| B | 3445 hCoV-19/Hungary/SRC-00572w/2020\|EPI_ISL_435431\|202 | ···················· | ···················· | ····················· | ···················· | ···················· | ···················· |
| B | 3446 hCoV-19/USA/AK-PHL151/2020\|EPI_ISL_435441\|2020-04- | ···················· | ···················· | ····················· | ···················· | ···················· | ···················· |
| B | 3447 hCoV-19/USA/AK-PHL153/2020\|EPI_ISL_435442\|2020-04- | ···················· | ···················· | ····················· | ···················· | ···················· | ···················· |
| B | 3448 hCoV-19/USA/AK-PHL180/2020\|EPI_ISL_435443\|2020-04- | ···················· | ···················· | ····················· | ···················· | ···················· | ···················· |
| B | 3449 hCoV-19/USA/AK-PHL193/2020\|EPI_ISL_435444\|2020-04- | ···················· | ···················· | ····················· | ···················· | ···················· | ···················· |
| B | 3450 hCoV-19/USA/LA-SR0140/2020\|EPI_ISL_435445\|2020-03- | ···················· | ···················· | ····················· | ···················· | ···················· | ···················· |
| B | 3451 hCoV-19/USA/LA-SR0141/2020\|EPI_ISL_435446\|2020-03- | ···················· | ···················· | ····················· | ···················· | ···················· | ···················· |
| B | 3452 hCoV-19/USA/LA-SR0143/2020\|EPI_ISL_435447\|2020-03- | ···················· | ···················· | ····················· | ···················· | ···················· | ···················· |
| B | 3453 hCoV-19/USA/LA-SR0144/2020\|EPI_ISL_435448\|2020-03- | ···················· | ···················· | ····················· | ···················· | ···················· | ···················· |
| B | 3454 hCoV-19/USA/LA-SR0145/2020\|EPI_ISL_435449\|2020-03- | ···················· | ···················· | ····················· | ···················· | ···················· | ···················· |
| B | 3455 hCoV-19/USA/LA-SR0146/2020\|EPI_ISL_435450\|2020-03- | ···················· | ···················· | ····················· | ···················· | ···················· | ···················· |
| B | 3456 hCoV-19/USA/LA-SR0148/2020\|EPI_ISL_435451\|2020-03- | ···················· | ···················· | ····················· | ···················· | ···················· | ···················· |
| B | 3457 hCoV-19/USA/LA-SR0149/2020\|EPI_ISL_435452\|2020-03- | ···················· | ···················· | ····················· | ···················· | ···················· | ···················· |
| B | 3458 hCoV-19/USA/LA-SR0150/2020\|EPI_ISL_435453\|2020-03- | ···················· | ···················· | ····················· | ···················· | ···················· | ···················· |
| B | 3459 hCoV-19/USA/LA-SR0151/2020\|EPI_ISL_435454\|2020-04- | ···················· | ···················· | ····················· | ···················· | ···················· | ···················· |
| B | 3460 hCoV-19/USA/LA-SR0153/2020\|EPI_ISL_435456\|2020-04- | ···················· | ···················· | ····················· | ···················· | ···················· | ···················· |
| B | 3461 hCoV-19/USA/LA-SR0154/2020\|EPI_ISL_435457\|2020-04- | ···················· | ···················· | ····················· | ···················· | ···················· | ···················· |
| B | 3462 hCoV-19/USA/LA-SR0155/2020\|EPI_ISL_435458\|2020-04- | ···················· | ···················· | ····················· | ···················· | ···················· | ···················· |
| B | 3463 hCoV-19/USA/LA-SR0156/2020\|EPI_ISL_435459\|2020-04- | ···················· | ···················· | ····················· | ···················· | ···················· | ···················· |
| B | 3464 hCoV-19/USA/LA-SR0157/2020\|EPI_ISL_435460\|2020-04- | ···················· | ···················· | ····················· | ···················· | ···················· | ···················· |
| B | 3465 hCoV-19/USA/LA-SR0158/2020\|EPI_ISL_435461\|2020-04- | ···················· | ···················· | ····················· | ···················· | ···················· | ···················· |
| B | 3466 hCoV-19/USA/LA-SR0159/2020\|EPI_ISL_435462\|2020-04- | ···················· | ···················· | ····················· | ···················· | ···················· | ···················· |
| B | 3467 hCoV-19/USA/LA-SR0161/2020\|EPI_ISL_435464\|2020-03- | ···················· | ···················· | ····················· | ···················· | ···················· | ···················· |
| B | 3468 hCoV-19/USA/LA-SR0162/2020\|EPI_ISL_435465\|2020-03- | ···················· | ···················· | ····················· | ···················· | ···················· | ···················· |
| B | 3469 hCoV-19/USA/LA-SR0163/2020\|EPI_ISL_435466\|2020-03- | ···················· | ···················· | ····················· | ···················· | ···················· | ···················· |
| B | 3470 hCoV-19/USA/LA-SR0164/2020\|EPI_ISL_435467\|2020-03- | ···················· | ···················· | ····················· | ···················· | ···················· | ···················· |
| B | 3471 hCoV-19/USA/LA-SR0166/2020\|EPI_ISL_435468\|2020-04- | ···················· | ···················· | ····················· | ···················· | ···················· | ···················· |
| B | 3472 hCoV-19/USA/LA-SR0167/2020\|EPI_ISL_435469\|2020-04- | ···················· | ···················· | ····················· | ···················· | ···················· | ···················· |
| B | 3473 hCoV-19/Russia/Moscow-GCBL1/2020\|EPI_ISL_436715\|20 | ···················· | ···················· | ····················· | ···················· | ···················· | ···················· |
| B | 3474 hCoV-19/Russia/Moscow-GCBL2/2020\|EPI_ISL_436716\|20 | ···················· | ···················· | ····················· | ···················· | ···················· | ···················· |
| B | 3475 hCoV-19/Russia/Moscow-GCBL3/2020\|EPI_ISL_436717\|20 | ···················· | ···················· | ····················· | ···················· | ···················· | ···················· |
| B | 3476 hCoV-19/Italy/ABR-IZSGC-TE5473/2020\|EPI_ISL_436718 | ···················· | ···················· | ····················· | ···················· | ···················· | ···················· |
| B | 3477 hCoV-19/Italy/ABR-IZSGC-TE5541/2020\|EPI_ISL_436719 | ···················· | ···················· | ····················· | ···················· | ···················· | ···················· |
| B | 3478 hCoV-19/Italy/ABR-IZSGC-TE5543/2020\|EPI_ISL_436720 | ···················· | ···················· | ····················· | ···················· | ···················· | ···················· |
| B | 3479 hCoV-19/Italy/ABR-IZSGC-TE5545/2020\|EPI_ISL_436721 | ···················· | ···················· | ····················· | ···················· | ···················· | ···················· |
| B | 3480 hCoV-19/Italy/ABR-IZSGC-TE5547/2020\|EPI_ISL_436722 | ···················· | ···················· | ····················· | ···················· | ···················· | ···················· |
| B | 3481 hCoV-19/Italy/ABR-IZSGC-TE5687/2020\|EPI_ISL_436723 | ···················· | ···················· | ····················· | ···················· | ···················· | ···················· |
| B | 3482 hCoV-19/Italy/ABR-IZSGC-TE5827/2020\|EPI_ISL_436724 | ···················· | ···················· | ····················· | ···················· | ···················· | ···················· |
| B | 3483 hCoV-19/Italy/ABR-IZSGC-TE27020/2020\|EPI_ISL_43672 | ···················· | ···················· | ····················· | ···················· | ···················· | ···················· |
| B | 3484 hCoV-19/Italy/ABR-IZSGC-TE26533/2020\|EPI_ISL_43672 | ···················· | ···················· | ····················· | ···················· | ···················· | ···················· |
| B | 3485 hCoV-19/Italy/ABR-IZSGC-TE26539/2020\|EPI_ISL_43672 | ···················· | ···················· | ····················· | ···················· | ···················· | ···················· |
| B | 3486 hCoV-19/Italy/ABR-IZSGC-TE26643/2020\|EPI_ISL_43672 | ···················· | ···················· | ····················· | ···················· | ···················· | ···················· |
| B | 3487 hCoV-19/Italy/ABR-IZSGC-TE26969/2020\|EPI_ISL_43673 | ···················· | ···················· | ····················· | ···················· | ···················· | ···················· |
| B | 3488 hCoV-19/Italy/ABR-IZSGC-TE26423/2020\|EPI_ISL_43673 | ···················· | ···················· | ····················· | ···················· | ···················· | ···················· |
| B | 3489 hCoV-19/Italy/ABR-IZSGC-TE26425/2020\|EPI_ISL_43673 | ···················· | ···················· | ····················· | ···················· | ···················· | ···················· |
| B | 3490 hCoV-19/England/LOND-D38C2/2020\|EPI_ISL_441003\|202 | ···················· | ···················· | ····················· | ···················· | ···················· | ···················· |
| B | 3491 hCoV-19/England/LOND-D38D1/2020\|EPI_ISL_441004\|202 | ···················· | ···················· | ····················· | ···················· | ···················· | ···················· |
| B | 3492 hCoV-19/England/LOND-D38E0/2020\|EPI_ISL_441005\|202 | ···················· | ···················· | ····················· | ···················· | ···················· | ···················· |
| B | 3493 hCoV-19/England/LOND-D38FF/2020\|EPI_ISL_441006\|202 | ···················· | ···················· | ····················· | ···················· | ···················· | ···················· |
| B | 3494 hCoV-19/England/LOND-D390B/2020\|EPI_ISL_441007\|202 | ···················· | ···················· | ····················· | ···················· | ···················· | ···················· |
| B | 3495 hCoV-19/England/LOND-D391A/2020\|EPI_ISL_441008\|202 | ···················· | ···················· | ····················· | ···················· | ···················· | ···················· |
| B | 3496 hCoV-19/England/LOND-D3929/2020\|EPI_ISL_441009\|202 | ·T·················· | ···················· | ····················· | ···················· | ···················· | ···················· |
| B | 3497 hCoV-19/England/LOND-D3938/2020\|EPI_ISL_441010\|202 | ···················· | ···················· | ····················· | ···················· | ···················· | ···················· |
| B | 3498 hCoV-19/England/LOND-D3947/2020\|EPI_ISL_441011\|202 | ···················· | ···················· | ····················· | ···················· | ···················· | ···················· |
| B | 3499 hCoV-19/England/LOND-D3956/2020\|EPI_ISL_441012\|202 | ···················· | ···················· | ····················· | ···················· | ···················· | ···················· |
| B | 3500 hCoV-19/England/LOND-D3965/2020\|EPI_ISL_441013\|202 | ···················· | ···················· | ····················· | ···················· | ···················· | ···················· |
| B | 3501 hCoV-19/England/LOND-D3974/2020\|EPI_ISL_441014\|202 | ···················· | ···················· | ····················· | ···················· | ···················· | ···················· |
| B | 3502 hCoV-19/England/LOND-D3983/2020\|EPI_ISL_441015\|202 | ···················· | ···················· | ····················· | ···················· | ···················· | ···················· |
| B | 3503 hCoV-19/England/LOND-D3992/2020\|EPI_ISL_441016\|202 | ···················· | ···················· | ····················· | ···················· | ···················· | ···················· |
| B | 3504 hCoV-19/England/LOND-D39A1/2020\|EPI_ISL_441017\|202 | ···················· | ···················· | ····················· | ···················· | ···················· | ···················· |
| B | 3505 hCoV-19/England/LOND-D39B0/2020\|EPI_ISL_441018\|202 | ···················· | ···················· | ····················· | ···················· | ···················· | ···················· |
| B | 3506 hCoV-19/England/LOND-D39CF/2020\|EPI_ISL_441019\|202 | ···················· | ···················· | ····················· | ···················· | ···················· | ···················· |
| B | 3507 hCoV-19/England/LOND-D39DE/2020\|EPI_ISL_441020\|202 | ···················· | ·A·················· | ····················· | ···················· | ···················· | ···················· |
| B | 3508 hCoV-19/England/LOND-D39FC/2020\|EPI_ISL_441021\|202 | ···················· | ···················· | ····················· | ···················· | ···················· | ···················· |
| B | 3509 hCoV-19/England/LOND-D3A08/2020\|EPI_ISL_441022\|202 | ···················· | ···················· | ····················· | ···················· | ···················· | ···················· |
| B | 3510 hCoV-19/England/LOND-D3A17/2020\|EPI_ISL_441023\|202 | ···················· | ···················· | ····················· | ···················· | ···················· | ···················· |
| B | 3511 hCoV-19/England/LOND-D3A26/2020\|EPI_ISL_441024\|202 | ···················· | ···················· | ····················· | ···················· | ···················· | ···················· |
| B | 3512 hCoV-19/England/CAMB-7CC47/2020\|EPI_ISL_442320\|202 | ···················· | ···················· | ····················· | ···················· | ···················· | ···················· |
| B | 3513 hCoV-19/England/CAMB-7BA99/2020\|EPI_ISL_442321\|202 | ···················· | ···················· | ····················· | ···················· | ···················· | ···················· |
| B | 3514 hCoV-19/England/CAMB-7CE9C/2020\|EPI_ISL_442334\|202 | ···················· | ···················· | ····················· | ···················· | ···················· | ···················· |
| B | 3515 hCoV-19/England/CAMB-7C4B8/2020\|EPI_ISL_442335\|202 | ···················· | ···················· | ····················· | ···················· | ···················· | ···················· |
| B | 3516 hCoV-19/England/CAMB-7B9BA/2020\|EPI_ISL_442336\|202 | ···················· | ···················· | ····················· | ···················· | ···················· | ···················· |
| B | 3517 hCoV-19/England/CAMB-7D411/2020\|EPI_ISL_442337\|202 | ···················· | ···················· | ····················· | ···················· | ···················· | ···················· |
| B | 3518 hCoV-19/England/CAMB-7D10B/2020\|EPI_ISL_442338\|202 | ···················· | ···················· | ····················· | ···················· | ···················· | ···················· |
| B | 3519 hCoV-19/England/CAMB-7D183/2020\|EPI_ISL_442339\|202 | ···················· | ···················· | ····················· | ···················· | ···················· | ···················· |
| B | 3520 hCoV-19/England/CAMB-7D790/2020\|EPI_ISL_442340\|202 | ···················· | ···················· | ····················· | ···················· | ···················· | ···················· |
| B | 3521 hCoV-19/England/CAMB-7CC65/2020\|EPI_ISL_442341\|202 | ···················· | ···················· | ····················· | ···················· | ···················· | ···················· |

| | | | | | | | |
|---|---|---|---|---|---|---|---|
| B | 3522 hCoV-19/England/CAMB-7CEF6/2020\|EPI_ISL_442342\|202 | ···················· | ···················· | ···················· | ···················· | ···················· | ···················· |
| B | 3523 hCoV-19/England/CAMB-7B774/2020\|EPI_ISL_442343\|202 | ···················· | ···················· | ···················· | ···················· | ···················· | ···················· |
| B | 3524 hCoV-19/England/SHEF-C9DFD/2020\|EPI_ISL_442350\|202 | ···················· | ···················· | ···················· | ···················· | ···················· | ···················· |
| B | 3525 hCoV-19/England/SHEF-C9FBB/2020\|EPI_ISL_442353\|202 | ···················· | ···················· | ···················· | ···················· | ···················· | ···················· |
| B | 3526 hCoV-19/England/SHEF-C9FF7/2020\|EPI_ISL_442354\|202 | ···················· | ···················· | ···················· | ···················· | ···················· | ···················· |
| B | 3527 hCoV-19/England/CAMB-83357/2020\|EPI_ISL_443651\|202 | ···················· | ···················· | ···················· | ···················· | ···················· | ···················· |
| B | 3528 hCoV-19/England/CAMB-824DD/2020\|EPI_ISL_443653\|202 | ···················· | ···················· | ···················· | ···················· | ···················· | ···················· |
| B | 3529 hCoV-19/England/CAMB-84392/2020\|EPI_ISL_443667\|202 | ···················· | ···················· | ···················· | ···················· | ···················· | ···················· |
| B | 3530 hCoV-19/England/CAMB-845C9/2020\|EPI_ISL_443668\|202 | ···················· | ···················· | ···················· | ···················· | ···················· | ···················· |
| B | 3531 hCoV-19/England/CAMB-822F1/2020\|EPI_ISL_443670\|202 | ···················· | ···················· | ···················· | ···················· | ···················· | ···················· |
| B | 3532 hCoV-19/England/CAMB-82D69/2020\|EPI_ISL_443672\|202 | ···················· | ···················· | ···················· | ···················· | ···················· | ···················· |
| B | 3533 hCoV-19/England/CAMB-825F8/2020\|EPI_ISL_443673\|202 | ···················· | ···················· | ···················· | ···················· | ···················· | ···················· |
| B | 3534 hCoV-19/England/CAMB-8268C/2020\|EPI_ISL_443674\|202 | ···················· | ···················· | ···················· | ···················· | ···················· | ···················· |
| B | 3535 hCoV-19/England/CAMB-1AA2E3/2020\|EPI_ISL_443676\|20 | ···················· | ···················· | ···················· | ···················· | ···················· | ···················· |
| B | 3536 hCoV-19/England/CAMB-82E1B/2020\|EPI_ISL_443677\|202 | ···················· | ···················· | ···················· | ···················· | ···················· | ···················· |
| B | 3537 hCoV-19/England/CAMB-82D4B/2020\|EPI_ISL_443680\|202 | ···················· | ···················· | ···················· | ···················· | ···················· | ···················· |
| B | 3538 hCoV-19/England/CAMB-82385/2020\|EPI_ISL_443681\|202 | ···················· | ···················· | ···················· | ···················· | ···················· | ···················· |
| B | 3539 hCoV-19/England/CAMB-1AA2F2/2020\|EPI_ISL_443682\|20 | ···················· | ···················· | ···················· | ···················· | ···················· | ···················· |
| B | 3540 hCoV-19/England/CAMB-832A5/2020\|EPI_ISL_443685\|202 | ···················· | ···················· | ···················· | ···················· | ···················· | ···················· |
| B | 3541 hCoV-19/England/BRIS-1258DF/2020\|EPI_ISL_443686\|20 | ···················· | ···················· | ···················· | ···················· | ···················· | ···················· |
| B | 3542 hCoV-19/England/BRIS-124928/2020\|EPI_ISL_443901\|20 | ···················· | ···················· | ···················· | ···················· | ···················· | ···················· |
| B | 3543 hCoV-19/England/BRIS-125EDD/2020\|EPI_ISL_443902\|20 | ···················· | ···················· | ···················· | ···················· | ···················· | ···················· |
| B | 3544 hCoV-19/England/BRIS-1252B3/2020\|EPI_ISL_443904\|20 | ···················· | ···················· | ···················· | ···················· | ···················· | ···················· |
| B | 3545 hCoV-19/England/BRIS-125620/2020\|EPI_ISL_443906\|20 | ···················· | ···················· | ···················· | ···················· | ···················· | ···················· |
| B | 3546 hCoV-19/England/BRIS-125383/2020\|EPI_ISL_443907\|20 | ···················· | ···················· | ···················· | ···················· | ···················· | ···················· |
| B | 3547 hCoV-19/England/BRIS-12572D/2020\|EPI_ISL_443909\|20 | ···················· | ···················· | ···················· | ···················· | ···················· | ···················· |
| B | 3548 hCoV-19/England/BRIS-124B40/2020\|EPI_ISL_443910\|20 | ···················· | ···················· | ···················· | ···················· | ···················· | ···················· |
| B | 3549 hCoV-19/England/BRIS-124973/2020\|EPI_ISL_443911\|20 | ···················· | ···················· | ···················· | ···················· | ···················· | ···················· |
| B | 3550 hCoV-19/England/BRIS-12482B/2020\|EPI_ISL_443912\|20 | ···················· | ···················· | ···················· | ···················· | ···················· | ···················· |
| B | 3551 hCoV-19/England/BRIS-125101/2020\|EPI_ISL_443914\|20 | ···················· | ···················· | ···················· | ···················· | ···················· | ···················· |
| B | 3552 hCoV-19/England/BRIS-125A33/2020\|EPI_ISL_443915\|20 | ···················· | ···················· | ···················· | ···················· | ···················· | ···················· |
| B | 3553 hCoV-19/England/BRIS-125B4F/2020\|EPI_ISL_443916\|20 | ···················· | ···················· | ···················· | ···················· | ···················· | ···················· |
| B | 3554 hCoV-19/England/BRIS-124982/2020\|EPI_ISL_443918\|20 | ···················· | ···················· | ···················· | ···················· | ···················· | ···················· |
| B | 3555 hCoV-19/England/BRIS-124BF5/2020\|EPI_ISL_443919\|20 | ···················· | ···················· | ···················· | ···················· | ···················· | ···················· |
| B | 3556 hCoV-19/Spain/CT-HUVH-VH4964/2020\|EPI_ISL_444980\|2 | ···················· | ···················· | ···················· | ···················· | ···················· | ···················· |
| B | 3557 hCoV-19/Spain/CT-HUVH-VH5081/2020\|EPI_ISL_444981\|2 | ···················· | ···················· | ···················· | ···················· | ···················· | ···················· |
| B | 3558 hCoV-19/Spain/CT-HUVH-VH5325/2020\|EPI_ISL_444982\|2 | ···················· | ···················· | ···················· | ···················· | ···················· | ···················· |
| B | 3559 hCoV-19/Spain/CT-HUVH-VH5994/2020\|EPI_ISL_444983\|2 | ···················· | ···················· | ···················· | ···················· | ···················· | ···················· |
| B | 3560 hCoV-19/Spain/CT-HUVH-VH6552/2020\|EPI_ISL_444984\|2 | ···················· | ···················· | ···················· | ···················· | ···················· | ···················· |
| B | 3561 hCoV-19/USA/SC_NHRC_02/2020\|EPI_ISL_444996\|2020-03 | ···················· | ···················· | ···················· | ···················· | ···················· | ···················· |
| B | 3562 hCoV-19/USA/SC_NHRC_03/2020\|EPI_ISL_444997\|2020-03 | ···················· | ···················· | ···················· | ···················· | ···················· | ···················· |
| B | 3563 hCoV-19/Guam/GU_NHG_01/2020\|EPI_ISL_444998\|2020-03 | ···················· | ···················· | ···················· | ···················· | ···················· | ···················· |
| B | 3564 hCoV-19/Guam/GU_NHG_02/2020\|EPI_ISL_444999\|2020-03 | ···················· | ···················· | ···················· | ···················· | ···················· | ···················· |
| B | 3565 hCoV-19/Wales/PHWC-2EA2C/2020\|EPI_ISL_446101\|2020- | ···················· | ···················· | ···················· | ···················· | ···················· | ···················· |
| B | 3566 hCoV-19/Wales/PHWC-2EA3B/2020\|EPI_ISL_446102\|2020- | ···················· | ···················· | ···················· | ···················· | ···················· | ···················· |
| B | 3567 hCoV-19/Wales/PHWC-2EA4A/2020\|EPI_ISL_446103\|2020- | ···················· | ···················· | ···················· | ···················· | ···················· | ···················· |
| B | 3568 hCoV-19/Wales/PHWC-2EB0B/2020\|EPI_ISL_446115\|2020- | ···················· | ···················· | ···················· | ···················· | ···················· | ···················· |
| B | 3569 hCoV-19/Wales/PHWC-2EE20/2020\|EPI_ISL_446160\|2020- | ···················· | ···················· | ···················· | ···················· | ···················· | ···················· |
| B | 3570 hCoV-19/Wales/PHWC-2EE4E/2020\|EPI_ISL_446162\|2020- | ···················· | ···················· | ···················· | ···················· | ···················· | ···················· |
| B | 3571 hCoV-19/Wales/PHWC-2EE6C/2020\|EPI_ISL_446164\|2020- | ···················· | ···················· | ···················· | ···················· | ···················· | ···················· |
| B | 3572 hCoV-19/Wales/PHWC-2EE99/2020\|EPI_ISL_446167\|2020- | ···················· | ···················· | ···················· | ···················· | ···················· | ···················· |
| B | 3573 hCoV-19/Wales/PHWC-2EED5/2020\|EPI_ISL_446171\|2020- | ···················· | ···················· | ···················· | ···················· | ···················· | ···················· |
| B | 3574 hCoV-19/Wales/PHWC-2EEE4/2020\|EPI_ISL_446172\|2020- | ···················· | ···················· | ···················· | ···················· | ···················· | ···················· |
| B | 3575 hCoV-19/Wales/PHWC-2EF3C/2020\|EPI_ISL_446177\|2020- | ···················· | ···················· | ···················· | ···················· | ···················· | ···················· |
| B | 3576 hCoV-19/Wales/PHWC-2EFA5/2020\|EPI_ISL_446182\|2020- | ···················· | ···················· | ···················· | ···················· | ···················· | ···················· |
| B | 3577 hCoV-19/Wales/PHWC-2EFB4/2020\|EPI_ISL_446183\|2020- | ···················· | ····A··············· | ···················· | ···················· | ···················· | ···················· |
| B | 3578 hCoV-19/Wales/PHWC-2EFD2/2020\|EPI_ISL_446184\|2020- | ···················· | ···················· | ···················· | ···················· | ···················· | ···················· |
| B | 3579 hCoV-19/Wales/PHWC-2EFF0/2020\|EPI_ISL_446186\|2020- | ···················· | ···················· | ···················· | ···················· | ···················· | ···················· |
| B | 3580 hCoV-19/Wales/PHWC-2F00B/2020\|EPI_ISL_446187\|2020- | ···················· | ···················· | ···················· | ···················· | ···················· | ···················· |
| B | 3581 hCoV-19/Wales/PHWC-2F038/2020\|EPI_ISL_446190\|2020- | ···················· | ···················· | ···················· | ···················· | ···················· | ···················· |
| B | 3582 hCoV-19/Wales/PHWC-2F056/2020\|EPI_ISL_446192\|2020- | ···················· | ···················· | ···················· | ···················· | ···················· | ···················· |
| B | 3583 hCoV-19/Wales/PHWC-2F065/2020\|EPI_ISL_446193\|2020- | ···················· | ···················· | ···················· | ···················· | ···················· | ···················· |
| B | 3584 hCoV-19/Wales/PHWC-2F0DE/2020\|EPI_ISL_446199\|2020- | ···················· | ···················· | ···················· | ···················· | ···················· | ···················· |
| B | 3585 hCoV-19/USA/NY-NYUMC296/2020\|EPI_ISL_430323\|2020-0 | ···················· | ···················· | ···················· | ···················· | ···················· | ···················· |
| B | 3586 hCoV-19/USA/NY-NYUMC297/2020\|EPI_ISL_430324\|2020-0 | ···················· | ···················· | ···················· | ···················· | ···················· | ···················· |
| B | 3587 hCoV-19/USA/NY-NYUMC298/2020\|EPI_ISL_430325\|2020-0 | ···················· | ···················· | ···················· | ···················· | ···················· | ···················· |
| B | 3588 hCoV-19/USA/NY-NYUMC299/2020\|EPI_ISL_430326\|2020-0 | ···················· | ···················· | ···················· | ···················· | ···················· | ···················· |
| B | 3589 hCoV-19/USA/NY-NYUMC300/2020\|EPI_ISL_430327\|2020-0 | ···················· | ···················· | ···················· | ···················· | ···················· | ···················· |
| B | 3590 hCoV-19/USA/NY-NYUMC301/2020\|EPI_ISL_430328\|2020-0 | ···················· | ···················· | ···················· | ···················· | ···················· | ···················· |
| B | 3591 hCoV-19/USA/NY-NYUMC302/2020\|EPI_ISL_430329\|2020-0 | ···················· | ···················· | ···················· | ···················· | ···················· | ···················· |
| B | 3592 hCoV-19/USA/NY-NYUMC303/2020\|EPI_ISL_430330\|2020-0 | ···················· | ···················· | ···················· | ···················· | ···················· | ···················· |
| B | 3593 hCoV-19/USA/NY-NYUMC304/2020\|EPI_ISL_430331\|2020-0 | ···················· | ···················· | ···················· | ···················· | ···················· | ···················· |
| B | 3594 hCoV-19/USA/NY-NYUMC305/2020\|EPI_ISL_430332\|2020-0 | ···················· | ···················· | ···················· | ···················· | ···················· | ···················· |
| B | 3595 hCoV-19/USA/NY-NYUMC306/2020\|EPI_ISL_430333\|2020-0 | ···················· | ···················· | ···················· | ···················· | ···················· | ···················· |
| B | 3596 hCoV-19/USA/NY-NYUMC307/2020\|EPI_ISL_430334\|2020-0 | ···················· | ···················· | ···················· | ···················· | ···················· | ···················· |
| B | 3597 hCoV-19/USA/NY-NYUMC310/2020\|EPI_ISL_430337\|2020-0 | ···················· | ···················· | ···················· | ···················· | ···················· | ···················· |
| B | 3598 hCoV-19/USA/NY-NYUMC311/2020\|EPI_ISL_430338\|2020-0 | ···················· | ···················· | ···················· | ···················· | ···················· | ···················· |
| B | 3599 hCoV-19/USA/NY-NYUMC312/2020\|EPI_ISL_430339\|2020-0 | ···················· | ···················· | ···················· | ···················· | ···················· | ···················· |

| | | | | | | | |
|---|---|---|---|---|---|---|---|
| B | 3600 hCoV-19/USA/NY-NYUMC313/2020\|EPI_ISL_430340\|2020-0 | .................... | .................... | .................... | .................... | .................... | .................... |
| B | 3601 hCoV-19/USA/NY-NYUMC314/2020\|EPI_ISL_430341\|2020-0 | .................... | .................... | .................... | .................... | .................... | .................... |
| B | 3602 hCoV-19/USA/NY-NYUMC315/2020\|EPI_ISL_430342\|2020-0 | .................... | .................... | .................... | .................... | .................... | .................... |
| B | 3603 hCoV-19/USA/NY-NYUMC316/2020\|EPI_ISL_430343\|2020-0 | .................... | .................... | .................... | .................... | .................... | .................... |
| B | 3604 hCoV-19/USA/NY-NYUMC317/2020\|EPI_ISL_430344\|2020-0 | .................... | .................... | .................... | .................... | .................... | .................... |
| B | 3605 hCoV-19/USA/NY-NYUMC318/2020\|EPI_ISL_430345\|2020-0 | .................... | .................... | .................... | .................... | .................... | .................... |
| B | 3606 hCoV-19/USA/NY-NYUMC319/2020\|EPI_ISL_430346\|2020-0 | .................... | .................... | .................... | .................... | .................... | .................... |
| B | 3607 hCoV-19/USA/NY-NYUMC320/2020\|EPI_ISL_430347\|2020-0 | .................... | .................... | .................... | .................... | .................... | .................... |
| B | 3608 hCoV-19/USA/NY-NYUMC321/2020\|EPI_ISL_430348\|2020-0 | .................... | .................... | .................... | .................... | .................... | .................... |
| B | 3609 hCoV-19/USA/NY-NYUMC322/2020\|EPI_ISL_430349\|2020-0 | .................... | .................... | .................... | .................... | .................... | .................... |
| B | 3610 hCoV-19/USA/NY-NYUMC323/2020\|EPI_ISL_430350\|2020-0 | .................... | .................... | .................... | .................... | .................... | .................... |
| B | 3611 hCoV-19/USA/NY-NYUMC324/2020\|EPI_ISL_430351\|2020-0 | .................... | .................... | .................... | .................... | .................... | .................... |
| B | 3612 hCoV-19/USA/NY-NYUMC325/2020\|EPI_ISL_430352\|2020-0 | .................... | .................... | .................... | .................... | .................... | .................... |
| B | 3613 hCoV-19/USA/NY-NYUMC326/2020\|EPI_ISL_430353\|2020-0 | .................... | .................... | .................... | .................... | .................... | .................... |
| B | 3614 hCoV-19/USA/NY-NYUMC327/2020\|EPI_ISL_430354\|2020-0 | .................... | .................... | .................... | .................... | .................... | .................... |
| B | 3615 hCoV-19/USA/NY-NYUMC350/2020\|EPI_ISL_430377\|2020-0 | .................... | .................... | .................... | .................... | .................... | .................... |
| B | 3616 hCoV-19/USA/NY-NYUMC351/2020\|EPI_ISL_430378\|2020-0 | .................... | .................... | .................... | .................... | .................... | .................... |
| B | 3617 hCoV-19/USA/NY-NYUMC352/2020\|EPI_ISL_430379\|2020-0 | .................... | .................... | .................... | .................... | .................... | .................... |
| B | 3618 hCoV-19/USA/NY-NYUMC353/2020\|EPI_ISL_430380\|2020-0 | .................... | .................... | .................... | .................... | .................... | .................... |
| B | 3619 hCoV-19/USA/NY-NYUMC354/2020\|EPI_ISL_430381\|2020-0 | .................... | .................... | .................... | .................... | .................... | .................... |
| B | 3620 hCoV-19/USA/NY-NYUMC355/2020\|EPI_ISL_430382\|2020-0 | .................... | .................... | .................... | .................... | .................... | .................... |
| B | 3621 hCoV-19/USA/NY-NYUMC356/2020\|EPI_ISL_430383\|2020-0 | .................... | .................... | .................... | .................... | .................... | .................... |
| B | 3622 hCoV-19/USA/NY-NYUMC357/2020\|EPI_ISL_430384\|2020-0 | .................... | .................... | .................... | .................... | .................... | .................... |
| B | 3623 hCoV-19/USA/NY-NYUMC358/2020\|EPI_ISL_430385\|2020-0 | .................... | .................... | .................... | .................... | .................... | .................... |
| B | 3624 hCoV-19/USA/NY-NYUMC359/2020\|EPI_ISL_430386\|2020-0 | .................... | .................... | .................... | .................... | .................... | .................... |
| B | 3625 hCoV-19/USA/NY-NYUMC360/2020\|EPI_ISL_430387\|2020-0 | .................... | .................... | .................... | .................... | .................... | .................... |
| B | 3626 hCoV-19/USA/NY-NYUMC361/2020\|EPI_ISL_430388\|2020-0 | .................... | .................... | .................... | .................... | .................... | .................... |
| B | 3627 hCoV-19/USA/NY-NYUMC362/2020\|EPI_ISL_430389\|2020-0 | .................... | .................... | .................... | .................... | .................... | .................... |
| B | 3628 hCoV-19/USA/NY-NYUMC363/2020\|EPI_ISL_430390\|2020-0 | .................... | .................... | .................... | .................... | .................... | .................... |
| B | 3629 hCoV-19/USA/NY-NYUMC364/2020\|EPI_ISL_430391\|2020-0 | .................... | .................... | .................... | .................... | .................... | .................... |
| B | 3630 hCoV-19/USA/NY-NYUMC365/2020\|EPI_ISL_430392\|2020-0 | .................... | .................... | .................... | .................... | .................... | .................... |
| B | 3631 hCoV-19/USA/NY-NYUMC366/2020\|EPI_ISL_430393\|2020-0 | .................... | .................... | .................... | .................... | .................... | .................... |
| B | 3632 hCoV-19/USA/NJ-NYUMC367/2020\|EPI_ISL_430394\|2020-0 | .................... | .................... | .................... | .................... | .................... | .................... |
| B | 3633 hCoV-19/USA/NY-NYUMC368/2020\|EPI_ISL_430395\|2020-0 | .................... | .................... | .................... | .................... | .................... | .................... |
| B | 3634 hCoV-19/USA/NY-NYUMC369/2020\|EPI_ISL_430396\|2020-0 | .................... | .................... | .................... | .................... | .................... | .................... |
| B | 3635 hCoV-19/USA/NY-NYUMC370/2020\|EPI_ISL_430397\|2020-0 | .................... | .................... | .................... | .................... | .................... | .................... |
| B | 3636 hCoV-19/USA/NY-NYUMC371/2020\|EPI_ISL_430398\|2020-0 | .................... | .................... | .................... | .................... | .................... | .................... |
| B | 3637 hCoV-19/USA/NY-NYUMC372/2020\|EPI_ISL_430399\|2020-0 | .................... | .................... | .................... | .................... | .................... | .................... |
| B | 3638 hCoV-19/England/NOTT-10ED28/2020\|EPI_ISL_432916\|20 | .................... | .................... | .................... | .................... | .................... | .................... |
| B | 3639 hCoV-19/England/NOTT-10ED73/2020\|EPI_ISL_432921\|20 | .................... | .................... | .................... | .................... | .................... | .................... |
| B | 3640 hCoV-19/England/NOTT-10EDCE/2020\|EPI_ISL_432925\|20 | .................... | .................... | .................... | .................... | .................... | .................... |
| B | 3641 hCoV-19/England/NOTT-10EE16/2020\|EPI_ISL_432930\|20 | .................... | .................... | .................... | .................... | .................... | .................... |
| B | 3642 hCoV-19/England/NOTT-10EE8F/2020\|EPI_ISL_432935\|20 | .................... | .................... | .................... | .................... | .................... | .................... |
| B | 3643 hCoV-19/England/NOTT-10EEBC/2020\|EPI_ISL_432938\|20 | .................... | .................... | .................... | .................... | .................... | .................... |
| B | 3644 hCoV-19/England/NOTT-10EF13/2020\|EPI_ISL_432942\|20 | .................... | .................... | .................... | .................... | .................... | .................... |
| B | 3645 hCoV-19/England/NOTT-10EF31/2020\|EPI_ISL_432944\|20 | .................... | .................... | .................... | .................... | .................... | .................... |
| B | 3646 hCoV-19/England/NOTT-10F000/2020\|EPI_ISL_432955\|20 | .................... | .................... | .................... | .................... | .................... | .................... |
| B | 3647 hCoV-19/England/NOTT-10F01F/2020\|EPI_ISL_432956\|20 | .................... | .................... | .................... | .................... | .................... | .................... |
| B | 3648 hCoV-19/England/NOTT-10F237/2020\|EPI_ISL_432979\|20 | .................... | .................... | .................... | .................... | .................... | .................... |
| B | 3649 hCoV-19/England/NOTT-10F282/2020\|EPI_ISL_432984\|20 | .................... | .................... | .................... | .................... | .................... | .................... |
| B | 3650 hCoV-19/England/NOTT-10F2CE/2020\|EPI_ISL_432987\|20 | .................... | .................... | .................... | .................... | .................... | .................... |
| B | 3651 hCoV-19/England/NOTT-10F2DD/2020\|EPI_ISL_432988\|20 | .................... | .................... | .................... | .................... | .................... | .................... |
| B | 3652 hCoV-19/England/NOTT-10F2FB/2020\|EPI_ISL_432990\|20 | .................... | .................... | .................... | .................... | .................... | .................... |
| B | 3653 hCoV-19/England/NOTT-10F334/2020\|EPI_ISL_432992\|20 | .................... | .................... | .................... | .................... | .................... | .................... |
| B | 3654 hCoV-19/England/NOTT-10F343/2020\|EPI_ISL_432993\|20 | .................... | .................... | .................... | .................... | .................... | .................... |
| B | 3655 hCoV-19/England/NOTT-10F3CB/2020\|EPI_ISL_432998\|20 | .................... | .................... | .................... | .................... | .................... | .................... |
| B | 3656 hCoV-19/USA/VA-DCLS-0167/2020\|EPI_ISL_437800\|2020- | .................... | .................... | .................... | .................... | .................... | .................... |
| B | 3657 hCoV-19/USA/VA-DCLS-0168/2020\|EPI_ISL_437801\|2020- | .................... | .................... | .................... | .................... | .................... | .................... |
| B | 3658 hCoV-19/England/LOND-D3895/2020\|EPI_ISL_441000\|202 | .................... | .................... | .................... | .................... | .................... | .................... |
| B | 3659 hCoV-19/England/LOND-D38A4/2020\|EPI_ISL_441001\|202 | .................... | .................... | .................... | .................... | .................... | .................... |
| B | 3660 hCoV-19/England/LOND-D38B3/2020\|EPI_ISL_441002\|202 | .................... | .................... | .................... | .................... | .................... | .................... |
| B | 3661 hCoV-19/England/LOND-D3A35/2020\|EPI_ISL_441025\|202 | .................... | .................... | .................... | .................... | .................... | .................... |
| B | 3662 hCoV-19/England/LOND-D3A44/2020\|EPI_ISL_441026\|202 | .................... | .................... | .................... | .................... | .................... | .................... |
| B | 3663 hCoV-19/England/LOND-D3A53/2020\|EPI_ISL_441027\|202 | .................... | .................... | .................... | .................... | .................... | .................... |
| B | 3664 hCoV-19/England/LOND-D3A62/2020\|EPI_ISL_441028\|202 | .................... | .................... | .................... | .................... | .................... | .................... |
| B | 3665 hCoV-19/England/LOND-D3A71/2020\|EPI_ISL_441029\|202 | .................... | .................... | .................... | .................... | .................... | .................... |
| B | 3666 hCoV-19/England/LOND-D3A80/2020\|EPI_ISL_441030\|202 | .................... | .................... | .................... | .................... | .................... | .................... |
| B | 3667 hCoV-19/England/LOND-D3A9F/2020\|EPI_ISL_441031\|202 | .................... | .................... | .................... | .................... | .................... | .................... |
| B | 3668 hCoV-19/England/LOND-D3AAE/2020\|EPI_ISL_441032\|202 | .................... | .................... | .................... | .................... | .................... | .................... |
| B | 3669 hCoV-19/England/LOND-D3ABD/2020\|EPI_ISL_441033\|202 | .................... | .................... | .................... | .................... | .................... | .................... |
| B | 3670 hCoV-19/England/LOND-D3ACC/2020\|EPI_ISL_441034\|202 | .................... | .................... | .................... | .................... | .................... | .................... |
| B | 3671 hCoV-19/England/LOND-D3DE1/2020\|EPI_ISL_441035\|202 | .................... | .................... | .................... | .................... | .................... | .................... |
| B | 3672 hCoV-19/England/LOND-D3DF0/2020\|EPI_ISL_441036\|202 | .................... | .................... | .................... | .................... | .................... | .................... |
| B | 3673 hCoV-19/England/LOND-D3E0C/2020\|EPI_ISL_441037\|202 | .................... | .................... | .................... | .................... | .................... | .................... |
| B | 3674 hCoV-19/England/LOND-D3E1B/2020\|EPI_ISL_441038\|202 | .................... | .................... | .................... | .................... | .................... | .................... |
| B | 3675 hCoV-19/England/LOND-D3E39/2020\|EPI_ISL_441039\|202 | .................... | .................... | .................... | .................... | .................... | .................... |
| B | 3676 hCoV-19/England/LOND-D3E48/2020\|EPI_ISL_441040\|202 | .................... | .................... | .................... | .................... | .................... | .................... |
| B | 3677 hCoV-19/England/LOND-D3E57/2020\|EPI_ISL_441041\|202 | .................... | .................... | .................... | .................... | .................... | .................... |

| | | | | | | | | |
|---|---|---|---|---|---|---|---|---|
| B | 3678 | hCoV-19/England/LOND-D3E66/2020\|EPI_ISL_441042\|202 | .................... | .................... | .................... | .................... | .................... | .................... |
| B | 3679 | hCoV-19/England/LOND-D3E75/2020\|EPI_ISL_441043\|202 | .................... | .A.................. | .................... | .................... | .................... | .................... |
| B | 3680 | hCoV-19/England/LOND-D3EA2/2020\|EPI_ISL_441045\|202 | .................... | .................... | .................... | .................... | .................... | .................... |
| B | 3681 | hCoV-19/England/LOND-D3EB1/2020\|EPI_ISL_441046\|202 | .................... | .................... | .................... | .................... | .................... | .................... |
| B | 3682 | hCoV-19/England/LOND-D3EFD/2020\|EPI_ISL_441047\|202 | .................... | .................... | .................... | .................... | .................... | .................... |
| B | 3683 | hCoV-19/England/LOND-D3F90/2020\|EPI_ISL_441049\|202 | .................... | .................... | .................... | .................... | .................... | .................... |
| B | 3684 | hCoV-19/England/LOND-D3FAF/2020\|EPI_ISL_441050\|202 | .................... | .................... | .................... | .................... | .................... | .................... |
| B | 3685 | hCoV-19/England/LOND-D3FBE/2020\|EPI_ISL_441051\|202 | .................... | .................... | .................... | .................... | .................... | .................... |
| B | 3686 | hCoV-19/England/CAMB-79794/2020\|EPI_ISL_441053\|202 | .................... | .................... | .................... | .................... | .................... | .................... |
| B | 3687 | hCoV-19/England/CAMB-78B99/2020\|EPI_ISL_441054\|202 | .................... | .................... | .................... | .................... | .................... | .................... |
| B | 3688 | hCoV-19/England/CAMB-7B677/2020\|EPI_ISL_441055\|202 | .................... | .................... | .................... | .................... | .................... | .................... |
| B | 3689 | hCoV-19/England/CAMB-7A766/2020\|EPI_ISL_441057\|202 | .................... | .................... | .................... | .................... | .................... | .................... |
| B | 3690 | hCoV-19/England/CAMB-79239/2020\|EPI_ISL_441058\|202 | .................... | .................... | .................... | .................... | .................... | .................... |
| B | 3691 | hCoV-19/England/CAMB-793FA/2020\|EPI_ISL_441059\|202 | .................... | .................... | .................... | .................... | .................... | .................... |
| B | 3692 | hCoV-19/England/CAMB-78B3F/2020\|EPI_ISL_441062\|202 | .................... | .................... | .................... | .................... | .................... | .................... |
| B | 3693 | hCoV-19/England/CAMB-791F0/2020\|EPI_ISL_441063\|202 | .................... | .................... | .................... | .................... | .................... | .................... |
| B | 3694 | hCoV-19/England/CAMB-797A3/2020\|EPI_ISL_441064\|202 | .................... | .................... | .................... | .................... | .................... | .................... |
| B | 3695 | hCoV-19/England/CAMB-792C0/2020\|EPI_ISL_441066\|202 | .................... | .................... | .................... | .................... | .................... | .................... |
| B | 3696 | hCoV-19/England/CAMB-7A55D/2020\|EPI_ISL_441068\|202 | .................... | .................... | .................... | .................... | .................... | .................... |
| B | 3697 | hCoV-19/England/CAMB-7D36F/2020\|EPI_ISL_442301\|202 | .................... | .................... | .................... | .................... | .................... | .................... |
| B | 3698 | hCoV-19/England/CAMB-7C807/2020\|EPI_ISL_442302\|202 | .................... | .................... | .................... | .................... | .................... | .................... |
| B | 3699 | hCoV-19/England/CAMB-7CF5D/2020\|EPI_ISL_442305\|202 | .................... | .................... | .................... | .................... | .................... | .................... |
| B | 3700 | hCoV-19/England/CAMB-7CF3F/2020\|EPI_ISL_442306\|202 | .................... | .................... | .................... | .................... | .................... | .................... |
| B | 3701 | hCoV-19/England/CAMB-7D89D/2020\|EPI_ISL_442307\|202 | .................... | .................... | .................... | .................... | .................... | .................... |
| B | 3702 | hCoV-19/England/CAMB-7DD25/2020\|EPI_ISL_442308\|202 | .................... | .................... | .................... | .................... | .................... | .................... |
| B | 3703 | hCoV-19/England/CAMB-7CF20/2020\|EPI_ISL_442309\|202 | .................... | .................... | .................... | .................... | .................... | .................... |
| B | 3704 | hCoV-19/England/CAMB-7C46D/2020\|EPI_ISL_442310\|202 | .................... | .................... | .................... | .................... | .................... | .................... |
| B | 3705 | hCoV-19/England/CAMB-7B70B/2020\|EPI_ISL_442312\|202 | .................... | .................... | .................... | .................... | .................... | .................... |
| B | 3706 | hCoV-19/England/CAMB-7CC29/2020\|EPI_ISL_442313\|202 | .................... | .................... | .................... | .................... | .................... | .................... |
| B | 3707 | hCoV-19/England/CAMB-7BA7B/2020\|EPI_ISL_442315\|202 | .................... | .................... | .................... | .................... | .................... | .................... |
| B | 3708 | hCoV-19/England/CAMB-7D1A1/2020\|EPI_ISL_442316\|202 | .................... | .................... | .................... | .................... | .................... | .................... |
| B | 3709 | hCoV-19/England/CAMB-7C5A6/2020\|EPI_ISL_442317\|202 | .................... | .................... | .................... | .................... | .................... | .................... |
| B | 3710 | hCoV-19/England/CAMB-7CD26/2020\|EPI_ISL_442318\|202 | .................... | .................... | .................... | .................... | .................... | .................... |
| B | 3711 | hCoV-19/England/CAMB-7CC0B/2020\|EPI_ISL_442319\|202 | .................... | .................... | .................... | .................... | .................... | .................... |
| B | 3712 | hCoV-19/England/CAMB-7DC82/2020\|EPI_ISL_442323\|202 | .................... | .................... | .................... | .................... | .................... | .................... |
| B | 3713 | hCoV-19/England/CAMB-7C421/2020\|EPI_ISL_442324\|202 | .................... | .................... | .................... | .................... | .................... | .................... |
| B | 3714 | hCoV-19/England/CAMB-7C403/2020\|EPI_ISL_442325\|202 | .................... | .................... | .................... | .................... | .................... | .................... |
| B | 3715 | hCoV-19/England/CAMB-7D754/2020\|EPI_ISL_442326\|202 | .................... | .................... | .................... | .................... | .................... | .................... |
| B | 3716 | hCoV-19/England/CAMB-7DC28/2020\|EPI_ISL_442327\|202 | .................... | .................... | .................... | .................... | .................... | .................... |
| B | 3717 | hCoV-19/England/CAMB-7CE8D/2020\|EPI_ISL_442329\|202 | .................... | .................... | .................... | .................... | .................... | .................... |
| B | 3718 | hCoV-19/England/CAMB-7C694/2020\|EPI_ISL_442330\|202 | .................... | .................... | .................... | .................... | .................... | .................... |
| B | 3719 | hCoV-19/England/CAMB-7B7FC/2020\|EPI_ISL_442331\|202 | .................... | .................... | .................... | .................... | .................... | .................... |
| B | 3720 | hCoV-19/England/CAMB-7D815/2020\|EPI_ISL_442332\|202 | .................... | .................... | .................... | .................... | .................... | .................... |
| B | 3721 | hCoV-19/England/CAMB-7D0D1/2020\|EPI_ISL_442333\|202 | .................... | .................... | .................... | .................... | .................... | .................... |
| B | 3722 | hCoV-19/England/SHEF-CA284/2020\|EPI_ISL_442357\|202 | .................... | .................... | .................... | .................... | .................... | .................... |
| B | 3723 | hCoV-19/England/SHEF-CA345/2020\|EPI_ISL_442358\|202 | .................... | .................... | .................... | .................... | .................... | .................... |
| B | 3724 | hCoV-19/England/SHEF-CAD1A/2020\|EPI_ISL_442370\|202 | .................... | .................... | .................... | .................... | .................... | .................... |
| B | 3725 | hCoV-19/England/SHEF-CAE08/2020\|EPI_ISL_442372\|202 | .................... | .................... | .................... | .................... | .................... | .................... |
| B | 3726 | hCoV-19/England/SHEF-CB1E0/2020\|EPI_ISL_442380\|202 | .................... | .................... | .................... | .................... | .................... | .................... |
| B | 3727 | hCoV-19/England/SHEF-CB61E/2020\|EPI_ISL_442387\|202 | .................... | .................... | .................... | .................... | .................... | .................... |
| B | 3728 | hCoV-19/England/SHEF-CB687/2020\|EPI_ISL_442390\|202 | .................... | .................... | .................... | .................... | .................... | .................... |
| B | 3729 | hCoV-19/England/SHEF-CBB2E/2020\|EPI_ISL_442397\|202 | .................... | .................... | .................... | .................... | .................... | .................... |
| B | 3730 | hCoV-19/England/SHEF-CBB5B/2020\|EPI_ISL_442398\|202 | .................... | .................... | .................... | .................... | .................... | .................... |
| B | 3731 | hCoV-19/England/CAMB-842B3/2020\|EPI_ISL_443621\|202 | .................... | .................... | .................... | .................... | .................... | .................... |
| B | 3732 | hCoV-19/England/CAMB-8316C/2020\|EPI_ISL_443622\|202 | .................... | .................... | .................... | .................... | .................... | .................... |
| B | 3733 | hCoV-19/England/CAMB-84338/2020\|EPI_ISL_443623\|202 | .................... | .................... | .................... | .................... | .................... | .................... |
| B | 3734 | hCoV-19/England/CAMB-82E2A/2020\|EPI_ISL_443625\|202 | .................... | .................... | .................... | .................... | .................... | .................... |
| B | 3735 | hCoV-19/England/CAMB-82464/2020\|EPI_ISL_443626\|202 | .................... | .................... | .................... | .................... | .................... | .................... |
| B | 3736 | hCoV-19/England/CAMB-8265F/2020\|EPI_ISL_443628\|202 | .................... | .................... | .................... | .................... | .................... | .................... |
| B | 3737 | hCoV-19/England/CAMB-1AA2B6/2020\|EPI_ISL_443629\|20 | .................... | .................... | .................... | .................... | .................... | .................... |
| B | 3738 | hCoV-19/England/CAMB-827B6/2020\|EPI_ISL_443630\|202 | .................... | .................... | .................... | .................... | .................... | .................... |
| B | 3739 | hCoV-19/England/CAMB-82938/2020\|EPI_ISL_443631\|202 | .................... | .................... | .................... | .................... | .................... | .................... |
| B | 3740 | hCoV-19/England/CAMB-84435/2020\|EPI_ISL_443632\|202 | .................... | .................... | .................... | .................... | .................... | .................... |
| B | 3741 | hCoV-19/England/CAMB-8423B/2020\|EPI_ISL_443633\|202 | .................... | .................... | .................... | .................... | .................... | .................... |
| B | 3742 | hCoV-19/England/CAMB-8306F/2020\|EPI_ISL_443634\|202 | .................... | .................... | .................... | .................... | .................... | .................... |
| B | 3743 | hCoV-19/England/CAMB-8369A/2020\|EPI_ISL_443636\|202 | .................... | .................... | .................... | .................... | .................... | .................... |
| B | 3744 | hCoV-19/England/CAMB-8277A/2020\|EPI_ISL_443637\|202 | .................... | .................... | .................... | .................... | .................... | .................... |
| B | 3745 | hCoV-19/England/CAMB-844AE/2020\|EPI_ISL_443639\|202 | .................... | .................... | .................... | .................... | .................... | .................... |
| B | 3746 | hCoV-19/England/CAMB-82288/2020\|EPI_ISL_443640\|202 | .................... | .................... | .................... | .................... | .................... | .................... |
| B | 3747 | hCoV-19/England/CAMB-1AA3B3/2020\|EPI_ISL_443644\|20 | .................... | .................... | .................... | .................... | .................... | .................... |
| B | 3748 | hCoV-19/England/CAMB-82877/2020\|EPI_ISL_443646\|202 | .................... | .................... | .................... | .................... | .................... | .................... |
| B | 3749 | hCoV-19/England/CAMB-84602/2020\|EPI_ISL_443648\|202 | .................... | .................... | .................... | .................... | .................... | .................... |
| B | 3750 | hCoV-19/England/CAMB-1AA526/2020\|EPI_ISL_443649\|20 | .................... | .................... | .................... | .................... | .................... | .................... |
| B | 3751 | hCoV-19/England/CAMB-830BA/2020\|EPI_ISL_443654\|202 | .................... | .................... | .................... | .................... | .................... | .................... |
| B | 3752 | hCoV-19/England/CAMB-1AA30E/2020\|EPI_ISL_443656\|20 | .................... | .A.................. | .................... | .................... | .................... | .................... |
| B | 3753 | hCoV-19/England/CAMB-82710/2020\|EPI_ISL_443657\|202 | .................... | .................... | .................... | .................... | .................... | .................... |
| B | 3754 | hCoV-19/England/CAMB-845AB/2020\|EPI_ISL_443658\|202 | .................... | .................... | .................... | .................... | .................... | .................... |
| B | 3755 | hCoV-19/England/CAMB-825E9/2020\|EPI_ISL_443661\|202 | .................... | .................... | .................... | .................... | .................... | .................... |

| | | | | | | | | |
|---|---|---|---|---|---|---|---|---|
| B | 3756 | hCoV-19/England/CAMB-1AA377/2020\|EPI_ISL_443662\|20 | ···················· | ···················· | ···················· | ···················· | ···················· | ···················· |
| B | 3757 | hCoV-19/England/CAMB-1AA368/2020\|EPI_ISL_443663\|20 | ···················· | ···················· | ···················· | ···················· | ···················· | ···················· |
| B | 3758 | hCoV-19/England/BRIS-12563F/2020\|EPI_ISL_443687\|20 | ···················· | ···················· | ···················· | ···················· | ···················· | ···················· |
| B | 3759 | hCoV-19/England/BRIS-125C00/2020\|EPI_ISL_443690\|20 | ···················· | ···················· | ···················· | ···················· | ···················· | ···················· |
| B | 3760 | hCoV-19/England/BRIS-125D1C/2020\|EPI_ISL_443691\|20 | ···················· | ···················· | ···················· | ···················· | ···················· | ···················· |
| B | 3761 | hCoV-19/England/BRIS-125453/2020\|EPI_ISL_443692\|20 | ···················· | ···················· | ···················· | ···················· | ···················· | ···················· |
| B | 3762 | hCoV-19/England/BRIS-1259CD/2020\|EPI_ISL_443697\|20 | ···················· | ···················· | ···················· | ···················· | ···················· | ···················· |
| B | 3763 | hCoV-19/England/BRIS-1252A4/2020\|EPI_ISL_443698\|20 | ···················· | ···················· | ···················· | ···················· | ···················· | ···················· |
| B | 3764 | hCoV-19/England/BRIS-125E28/2020\|EPI_ISL_443699\|20 | ···················· | ····A··············· | ···················· | ···················· | ···················· | ···················· |
| B | 3765 | hCoV-19/Denmark/ALAB-SSI-775/2020\|EPI_ISL_444900\|2 | ···················· | ···················· | ···················· | ···················· | ···················· | ···················· |
| B | 3766 | hCoV-19/Denmark/ALAB-SSI-779/2020\|EPI_ISL_444901\|2 | ···················· | ···················· | ···················· | ···················· | ···················· | ···················· |
| B | 3767 | hCoV-19/Denmark/ALAB-SSI-780/2020\|EPI_ISL_444902\|2 | ···················· | ···················· | ···················· | ···················· | ···················· | ···················· |
| B | 3768 | hCoV-19/Denmark/ALAB-SSI-782/2020\|EPI_ISL_444903\|2 | ···················· | ···················· | ···················· | ···················· | ···················· | ···················· |
| B | 3769 | hCoV-19/Denmark/ALAB-SSI-785/2020\|EPI_ISL_444904\|2 | ···················· | ···················· | ···················· | ···················· | ···················· | ···················· |
| B | 3770 | hCoV-19/Denmark/ALAB-SSI-787/2020\|EPI_ISL_444905\|2 | ···················· | ···················· | ···················· | ···················· | ···················· | ···················· |
| B | 3771 | hCoV-19/Denmark/ALAB-SSI-789/2020\|EPI_ISL_444906\|2 | ···················· | ···················· | ···················· | ···················· | ···················· | ···················· |
| B | 3772 | hCoV-19/Denmark/ALAB-SSI-793/2020\|EPI_ISL_444908\|2 | ···················· | ···················· | ···················· | ···················· | ···················· | ···················· |
| B | 3773 | hCoV-19/Denmark/ALAB-SSI-794/2020\|EPI_ISL_444909\|2 | ···················· | ···················· | ···················· | ···················· | ···················· | ···················· |
| B | 3774 | hCoV-19/Denmark/ALAB-SSI-795/2020\|EPI_ISL_444910\|2 | ···················· | ···················· | ···················· | ···················· | ···················· | ···················· |
| B | 3775 | hCoV-19/Denmark/ALAB-SSI-797/2020\|EPI_ISL_444911\|2 | ···················· | ···················· | ···················· | ···················· | ···················· | ···················· |
| B | 3776 | hCoV-19/Denmark/ALAB-SSI-799/2020\|EPI_ISL_444912\|2 | ···················· | ···················· | ···················· | ···················· | ···················· | ···················· |
| B | 3777 | hCoV-19/Denmark/ALAB-SSI-804/2020\|EPI_ISL_444915\|2 | ···················· | ···················· | ···················· | ···················· | ···················· | ···················· |
| B | 3778 | hCoV-19/Denmark/ALAB-SSI-805/2020\|EPI_ISL_444916\|2 | ···················· | ···················· | ···················· | ···················· | ···················· | ···················· |
| B | 3779 | hCoV-19/Denmark/ALAB-SSI-806/2020\|EPI_ISL_444917\|2 | ···················· | ···················· | ···················· | ···················· | ···················· | ···················· |
| B | 3780 | hCoV-19/Denmark/ALAB-SSI-809/2020\|EPI_ISL_444918\|2 | ···················· | ···················· | ···················· | ···················· | ···················· | ···················· |
| B | 3781 | hCoV-19/Denmark/ALAB-SSI-812/2020\|EPI_ISL_444920\|2 | ···················· | ···················· | ···················· | ···················· | ···················· | ···················· |
| B | 3782 | hCoV-19/Denmark/ALAB-SSI-815/2020\|EPI_ISL_444921\|2 | ···················· | ···················· | ···················· | ···················· | ···················· | ···················· |
| B | 3783 | hCoV-19/Denmark/ALAB-SSI-819/2020\|EPI_ISL_444922\|2 | ···················· | ···················· | ···················· | ···················· | ···················· | ···················· |
| B | 3784 | hCoV-19/Denmark/ALAB-SSI-821/2020\|EPI_ISL_444923\|2 | ···················· | ···················· | ···················· | ···················· | ···················· | ···················· |
| B | 3785 | hCoV-19/Denmark/ALAB-SSI-826/2020\|EPI_ISL_444924\|2 | ···················· | ···················· | ···················· | ···················· | ···················· | ···················· |
| B | 3786 | hCoV-19/Denmark/ALAB-SSI-827/2020\|EPI_ISL_444925\|2 | ···················· | ···················· | ···················· | ···················· | ···················· | ···················· |
| B | 3787 | hCoV-19/Denmark/ALAB-SSI-828/2020\|EPI_ISL_444926\|2 | ···················· | ···················· | ···················· | ···················· | ···················· | ···················· |
| B | 3788 | hCoV-19/Denmark/ALAB-SSI-831/2020\|EPI_ISL_444927\|2 | ···················· | ···················· | ···················· | ···················· | ···················· | ···················· |
| B | 3789 | hCoV-19/Denmark/ALAB-SSI-832/2020\|EPI_ISL_444928\|2 | ···················· | ···················· | ···················· | ···················· | ···················· | ···················· |
| B | 3790 | hCoV-19/Denmark/ALAB-SSI-833/2020\|EPI_ISL_444929\|2 | ···················· | ···················· | ···················· | ···················· | ···················· | ···················· |
| B | 3791 | hCoV-19/Denmark/ALAB-SSI-849/2020\|EPI_ISL_444940\|2 | ···················· | ···················· | ···················· | ···················· | ···················· | ···················· |
| B | 3792 | hCoV-19/Denmark/ALAB-SSI-876/2020\|EPI_ISL_444952\|2 | ···················· | ···················· | ···················· | ···················· | ···················· | ···················· |
| B | 3793 | hCoV-19/Denmark/ALAB-SSI-883/2020\|EPI_ISL_444954\|2 | ···················· | ···················· | ···················· | ···················· | ···················· | ···················· |
| B | 3794 | hCoV-19/Denmark/ALAB-SSI-885/2020\|EPI_ISL_444955\|2 | ···················· | ···················· | ···················· | ···················· | ···················· | ···················· |
| B | 3795 | hCoV-19/Denmark/ALAB-SSI-887/2020\|EPI_ISL_444956\|2 | ···················· | ···················· | ···················· | ···················· | ···················· | ···················· |
| B | 3796 | hCoV-19/Denmark/ALAB-SSI-888/2020\|EPI_ISL_444957\|2 | ···················· | ···················· | ···················· | ···················· | ···················· | ···················· |
| B | 3797 | hCoV-19/Denmark/ALAB-SSI-890/2020\|EPI_ISL_444958\|2 | ···················· | ···················· | ···················· | ···················· | ···················· | ···················· |
| B | 3798 | hCoV-19/Denmark/ALAB-SSI-892/2020\|EPI_ISL_444959\|2 | ···················· | ···················· | ···················· | ···················· | ···················· | ···················· |
| B | 3799 | hCoV-19/Denmark/ALAB-SSI-893/2020\|EPI_ISL_444960\|2 | ···················· | ···················· | ···················· | ···················· | ···················· | ···················· |
| B | 3800 | hCoV-19/Denmark/ALAB-SSI-894/2020\|EPI_ISL_444961\|2 | ···················· | ···················· | ···················· | ···················· | ···················· | ···················· |
| B | 3801 | hCoV-19/Denmark/ALAB-SSI-896/2020\|EPI_ISL_444963\|2 | ···················· | ···················· | ···················· | ···················· | ···················· | ···················· |
| B | 3802 | hCoV-19/Denmark/ALAB-SSI-899/2020\|EPI_ISL_444964\|2 | ···················· | ···················· | ···················· | ···················· | ···················· | ···················· |
| B | 3803 | hCoV-19/Denmark/ALAB-SSI-902/2020\|EPI_ISL_444965\|2 | ···················· | ···················· | ···················· | ···················· | ···················· | ···················· |
| B | 3804 | hCoV-19/Denmark/ALAB-SSI-904/2020\|EPI_ISL_444966\|2 | ···················· | ···················· | ···················· | ···················· | ···················· | ···················· |
| B | 3805 | hCoV-19/Denmark/ALAB-SSI-905/2020\|EPI_ISL_444967\|2 | ···················· | ···················· | ···················· | ···················· | ···················· | ···················· |
| B | 3806 | hCoV-19/Denmark/ALAB-SSI-907/2020\|EPI_ISL_444968\|2 | ···················· | ···················· | ···················· | ···················· | ···················· | ···················· |
| B | 3807 | hCoV-19/Guangdong/SYSU-IHV/2020\|EPI_ISL_444969\|202 | ···················· | ···················· | ···················· | ···················· | ···················· | ···················· |
| B | 3808 | hCoV-19/Spain/CT-HUVH-VH6372/2020\|EPI_ISL_444971\|2 | ···················· | ···················· | ···················· | ···················· | ···················· | ···················· |
| B | 3809 | hCoV-19/Spain/CT-HUVH-VH6819/2020\|EPI_ISL_444972\|2 | ···················· | ···················· | ···················· | ···················· | ···················· | ···················· |
| B | 3810 | hCoV-19/Spain/CT-HUVH-VH7280/2020\|EPI_ISL_444973\|2 | ···················· | ···················· | ···················· | ···················· | ···················· | ···················· |
| B | 3811 | hCoV-19/Spain/CT-HUVH-VH7458/2020\|EPI_ISL_444974\|2 | ···················· | ···················· | ···················· | ···················· | ···················· | ···················· |
| B | 3812 | hCoV-19/Spain/CT-HUVH-VH1407/2020\|EPI_ISL_444975\|2 | ···················· | ···················· | ···················· | ···················· | ···················· | ···················· |
| B | 3813 | hCoV-19/Spain/CT-HUVH-VH7773/2020\|EPI_ISL_444976\|2 | ···················· | ···················· | ···················· | ···················· | ···················· | ···················· |
| B | 3814 | hCoV-19/Spain/CT-HUVH-VH1438/2020\|EPI_ISL_444977\|2 | ···················· | ···················· | ···················· | ···················· | ···················· | ···················· |
| B | 3815 | hCoV-19/Spain/CT-HUVH-VH4141/2020\|EPI_ISL_444978\|2 | ···················· | ···················· | ···················· | ···················· | ···················· | ···················· |
| B | 3816 | hCoV-19/Spain/CT-HUVH-VH4488/2020\|EPI_ISL_444979\|2 | ···················· | ···················· | ···················· | ···················· | ···················· | ···················· |
| B | 3817 | hCoV-19/Spain/CT-HUVH-VH6837/2020\|EPI_ISL_444985\|2 | ···················· | ···················· | ···················· | ···················· | ···················· | ···················· |
| B | 3818 | hCoV-19/Spain/CT-HUVH-VH8938/2020\|EPI_ISL_444986\|2 | ···················· | ···················· | ···················· | ···················· | ···················· | ···················· |
| B | 3819 | hCoV-19/Spain/CT-HUVH-VH9434/2020\|EPI_ISL_444987\|2 | ···················· | ···················· | ···················· | ···················· | ···················· | ···················· |
| B | 3820 | hCoV-19/Spain/CT-HUVH-VH0851/2020\|EPI_ISL_444988\|2 | ···················· | ···················· | ···················· | ···················· | ···················· | ···················· |
| B | 3821 | hCoV-19/Spain/CT-HUVH-VH1484/2020\|EPI_ISL_444989\|2 | ···················· | ···················· | ···················· | ···················· | ···················· | ···················· |
| B | 3822 | hCoV-19/Spain/CT-HUVH-VH1713/2020\|EPI_ISL_444990\|2 | ···················· | ···················· | ···················· | ···················· | ···················· | ···················· |
| B | 3823 | hCoV-19/USA/CA-NHRC-01/2020\|EPI_ISL_444994\|2020-03 | ···················· | ···················· | ···················· | ···················· | ···················· | ···················· |
| B | 3824 | hCoV-19/USA/SC_NHRC_01/2020\|EPI_ISL_444995\|2020-03 | ···················· | ···················· | ···················· | ···················· | ···················· | ···················· |
| B | 3825 | hCoV-19/USA/MI-MDHHS-SC20201/2020\|EPI_ISL_447172\|2 | ···················· | ···················· | ···················· | ···················· | ···················· | ···················· |
| B | 3826 | hCoV-19/USA/MI-MDHHS-SC20202/2020\|EPI_ISL_447173\|2 | ···················· | ···················· | ···················· | ···················· | ···················· | ···················· |
| B | 3827 | hCoV-19/USA/MI-MDHHS-SC20203/2020\|EPI_ISL_447174\|2 | ···················· | ···················· | ···················· | ···················· | ···················· | ···················· |
| B | 3828 | hCoV-19/USA/MI-MDHHS-SC20204/2020\|EPI_ISL_447175\|2 | ···················· | ···················· | ···················· | ···················· | ···················· | ···················· |
| B | 3829 | hCoV-19/USA/MI-MDHHS-SC20206/2020\|EPI_ISL_447177\|2 | ···················· | ···················· | ···················· | ···················· | ···················· | ···················· |
| B | 3830 | hCoV-19/USA/MI-MDHHS-SC20207/2020\|EPI_ISL_447178\|2 | ···················· | ···················· | ···················· | ···················· | ···················· | ···················· |
| B | 3831 | hCoV-19/USA/MI-MDHHS-SC20208/2020\|EPI_ISL_447179\|2 | ···················· | ···················· | ···················· | ···················· | ···················· | ···················· |
| B | 3832 | hCoV-19/USA/MI-MDHHS-SC20212/2020\|EPI_ISL_447183\|2 | ···················· | ···················· | ···················· | ···················· | ···················· | ···················· |
| B | 3833 | hCoV-19/USA/MI-MDHHS-SC20213/2020\|EPI_ISL_447184\|2 | ···················· | ···················· | ···················· | ···················· | ···················· | ···················· |

| | | | | | | | | |
|---|---|---|---|---|---|---|---|---|
| B | 3834 | hCoV-19/USA/MI-MDHHS-SC20219/2020\|EPI_ISL_447189\|2 | .................... | .................... | ..................... | .................... | .................... | .................... |
| B | 3835 | hCoV-19/USA/MI-MDHHS-SC20220/2020\|EPI_ISL_447190\|2 | .................... | .................... | ..................... | .................... | .................... | .................... |
| B | 3836 | hCoV-19/USA/MI-MDHHS-SC20221/2020\|EPI_ISL_447191\|2 | .................... | .................... | ..................... | .................... | .................... | .................... |
| B | 3837 | hCoV-19/USA/MI-MDHHS-SC20223/2020\|EPI_ISL_447193\|2 | .................... | .................... | ..................... | .................... | .................... | .................... |
| B | 3838 | hCoV-19/USA/MI-MDHHS-SC20224/2020\|EPI_ISL_447194\|2 | .................... | .................... | ..................... | .................... | .................... | .................... |
| B | 3839 | hCoV-19/USA/MI-MDHHS-SC20226/2020\|EPI_ISL_447196\|2 | .................... | .................... | ..................... | .................... | .................... | .................... |
| B | 3840 | hCoV-19/USA/MI-MDHHS-SC20227/2020\|EPI_ISL_447197\|2 | .................... | .................... | ..................... | .................... | .................... | .................... |
| B | 3841 | hCoV-19/England/NOTT-10DF8D/2020\|EPI_ISL_432900\|20 | .................... | .................... | ..................... | .................... | .................... | .................... |
| B | 3842 | hCoV-19/England/NOTT-10EFB9/2020\|EPI_ISL_432951\|20 | .................... | .................... | ..................... | .................... | .................... | .................... |
| B | 3843 | hCoV-19/England/NOTT-10EFF5/2020\|EPI_ISL_432954\|20 | .................... | .................... | ..................... | .................... | .................... | .................... |
| B | 3844 | hCoV-19/England/NOTT-10F0B5/2020\|EPI_ISL_432961\|20 | .................... | .................... | ..................... | .................... | .................... | .................... |
| B | 3845 | hCoV-19/England/NOTT-10F0E2/2020\|EPI_ISL_432963\|20 | .................... | .................... | ..................... | .................... | .................... | .................... |
| B | 3846 | hCoV-19/England/NOTT-10F10D/2020\|EPI_ISL_432965\|20 | .................... | .................... | ..................... | .................... | .................... | .................... |
| B | 3847 | hCoV-19/England/NOTT-10F12B/2020\|EPI_ISL_432967\|20 | .................... | .................... | ..................... | .................... | .................... | .................... |
| B | 3848 | hCoV-19/England/NOTT-10F176/2020\|EPI_ISL_432970\|20 | .................... | .................... | ..................... | .................... | .................... | .................... |
| B | 3849 | hCoV-19/India/DL-NCDC-01457/2020\|EPI_ISL_435108\|20 | .................... | .................... | ..................... | .................... | .................... | .................... |
| B | 3850 | hCoV-19/India/DL-NCDC-01638/2020\|EPI_ISL_435109\|20 | .................... | .................... | ..................... | .................... | .................... | .................... |
| B | 3851 | hCoV-19/India/DL-NCDC-01501/2020\|EPI_ISL_435110\|20 | .................... | .................... | ..................... | .................... | ......N............. | .................... |
| B | 3852 | hCoV-19/India/DL-NCDC-02415/2020\|EPI_ISL_435111\|20 | .................... | .................... | ..................... | .................... | .................... | .................... |
| B | 3853 | hCoV-19/India/BR-NCDC-02251/2020\|EPI_ISL_435112\|20 | .................... | .................... | ..................... | .................... | .................... | .................... |
| B | 3854 | hCoV-19/DRC/1398/2020\|EPI_ISL_435114\|2020-04-07 | .................... | .................... | ..................... | .................... | .................... | .................... |
| B | 3855 | hCoV-19/United Arab Emirates/L0/2020\|EPI_ISL_43511 | .................... | .................... | ..................... | .................... | .................... | .................... |
| B | 3856 | hCoV-19/United Arab Emirates/L068/2020\|EPI_ISL_435 | .................... | .................... | ..................... | .................... | .................... | .................... |
| B | 3857 | hCoV-19/United Arab Emirates/L0184/2020\|EPI_ISL_43 | .................... | .................... | ..................... | .................... | .................... | .................... |
| B | 3858 | hCoV-19/United Arab Emirates/L0231/2020\|EPI_ISL_43 | .................... | .................... | ..................... | .................... | .................... | .................... |
| B | 3859 | hCoV-19/United Arab Emirates/L0484/2020\|EPI_ISL_43 | .................... | .................... | ..................... | .................... | .................... | .................... |
| B | 3860 | hCoV-19/United Arab Emirates/L0879/2020\|EPI_ISL_43 | .................... | .................... | ..................... | .................... | .................... | .................... |
| B | 3861 | hCoV-19/United Arab Emirates/L0881/2020\|EPI_ISL_43 | .................... | .................... | ..................... | .................... | .................... | .................... |
| B | 3862 | hCoV-19/United Arab Emirates/L0904/2020\|EPI_ISL_43 | .................... | .................... | ..................... | .................... | .................... | .................... |
| B | 3863 | hCoV-19/United Arab Emirates/L1014/2020\|EPI_ISL_43 | .................... | .................... | ..................... | .................... | .................... | .................... |
| B | 3864 | hCoV-19/United Arab Emirates/L1076/2020\|EPI_ISL_43 | .................... | .................... | ..................... | .................... | .................... | .................... |
| B | 3865 | hCoV-19/United Arab Emirates/L1758/2020\|EPI_ISL_43 | .................... | .................... | ..................... | .................... | .................... | .................... |
| B | 3866 | hCoV-19/United Arab Emirates/L2185/2020\|EPI_ISL_43 | .................... | .................... | ..................... | .................... | .................... | .................... |
| B | 3867 | hCoV-19/United Arab Emirates/L2409/2020\|EPI_ISL_43 | .................... | .................... | ..................... | .................... | .................... | .................... |
| B | 3868 | hCoV-19/United Arab Emirates/L3779/2020\|EPI_ISL_43 | .................... | .................... | ..................... | .................... | .................... | .................... |
| B | 3869 | hCoV-19/United Arab Emirates/L4184/2020\|EPI_ISL_43 | .................... | .................... | ..................... | .................... | .................... | .................... |
| B | 3870 | hCoV-19/United Arab Emirates/L4280/2020\|EPI_ISL_43 | .................... | .................... | ..................... | .................... | .................... | .................... |
| B | 3871 | hCoV-19/United Arab Emirates/L4682/2020\|EPI_ISL_43 | .................... | .................... | ..................... | .................... | .................... | .................... |
| B | 3872 | hCoV-19/United Arab Emirates/L5621/2020\|EPI_ISL_43 | .................... | .................... | ..................... | .................... | .................... | .................... |
| B | 3873 | hCoV-19/United Arab Emirates/L5630/2020\|EPI_ISL_43 | .................... | .................... | ..................... | .................... | .................... | .................... |
| B | 3874 | hCoV-19/United Arab Emirates/L6599/2020\|EPI_ISL_43 | .................... | .................... | ..................... | .................... | .................... | .................... |
| B | 3875 | hCoV-19/United Arab Emirates/L6627/2020\|EPI_ISL_43 | .................... | .................... | ..................... | .................... | .................... | .................... |
| B | 3876 | hCoV-19/United Arab Emirates/L7356/2020\|EPI_ISL_43 | .................... | .................... | ..................... | .................... | .................... | .................... |
| B | 3877 | hCoV-19/United Arab Emirates/L9440/2020\|EPI_ISL_43 | .................... | .................... | ..................... | .................... | .................... | .................... |
| B | 3878 | hCoV-19/United Arab Emirates/L9766/2020\|EPI_ISL_43 | .................... | .................... | ..................... | .................... | .................... | .................... |
| B | 3879 | hCoV-19/United Arab Emirates/L9768/2020\|EPI_ISL_43 | .................... | .................... | ..................... | .................... | .................... | .................... |
| B | 3880 | hCoV-19/Spain/MD-H12-LP28-2522/2020\|EPI_ISL_435144 | .................... | .................... | ..................... | .................... | .................... | .................... |
| B | 3881 | hCoV-19/Italy/ABR-IZSGC-TE6644/2020\|EPI_ISL_435145 | .................... | .................... | ..................... | .................... | .................... | .................... |
| B | 3882 | hCoV-19/Italy/ABR-IZSGC-TE12337/2020\|EPI_ISL_43514 | .................... | .................... | ..................... | .................... | .................... | .................... |
| B | 3883 | hCoV-19/Italy/ABR-IZSGC-TE12759/2020\|EPI_ISL_43514 | .................... | .................... | ..................... | .................... | .................... | .................... |
| B | 3884 | hCoV-19/Italy/ABR-IZSGC-TE13491/2020\|EPI_ISL_43514 | .................... | .................... | ..................... | .................... | .................... | .................... |
| B | 3885 | hCoV-19/Italy/ABR-IZSGC-TE13457/2020\|EPI_ISL_43514 | .................... | .................... | ..................... | .................... | .................... | .................... |
| B | 3886 | hCoV-19/Italy/ABR-IZSGC-TE12774/2020\|EPI_ISL_43515 | .................... | .................... | ..................... | .................... | .................... | .................... |
| B | 3887 | hCoV-19/Italy/ABR-IZSGC-TE13484/2020\|EPI_ISL_43515 | .................... | .................... | ..................... | .................... | .................... | .................... |
| B | 3888 | hCoV-19/Italy/ABR-IZSGC-TE13858/2020\|EPI_ISL_43515 | .................... | .................... | ..................... | .................... | .................... | .................... |
| B | 3889 | hCoV-19/Italy/ABR-IZSGC-TE14297/2020\|EPI_ISL_43515 | .................... | .................... | ..................... | .................... | .................... | .................... |
| B | 3890 | hCoV-19/Italy/ABR-IZSGC-TE14162/2020\|EPI_ISL_43515 | .................... | .................... | ..................... | .................... | .................... | .................... |
| B | 3891 | hCoV-19/Italy/ABR-IZSGC-TE14168/2020\|EPI_ISL_43515 | .................... | .................... | ..................... | .................... | .................... | .................... |
| B | 3892 | hCoV-19/DRC/1715/2020\|EPI_ISL_435156\|2020-04-10 | .................... | .................... | ..................... | .................... | .................... | .................... |
| B | 3893 | hCoV-19/DRC/2120/2020\|EPI_ISL_435163\|2020-04-12 | .................... | .................... | ..................... | .................... | .................... | .................... |
| B | 3894 | hCoV-19/India/BR-NCDC-2515/2020\|EPI_ISL_436417\|202 | .................... | .................... | ..................... | .................... | .................... | .................... |
| B | 3895 | hCoV-19/India/TN-NCDC-2517/2020\|EPI_ISL_436418\|202 | .................... | .................... | ..................... | .................... | .................... | .................... |
| B | 3896 | hCoV-19/India/BR-NCDC-2519/2020\|EPI_ISL_436419\|202 | .................... | .................... | ..................... | .................... | .................... | .................... |
| B | 3897 | hCoV-19/India/DL-NCDC-3170/2020\|EPI_ISL_436425\|202 | .................... | .................... | ..................... | .................... | .................... | .................... |
| B | 3898 | hCoV-19/India/AP-NCDC-3936/2020\|EPI_ISL_436440\|202 | .................... | .................... | ..................... | .................... | ..........M......... | .................... |
| B | 3899 | hCoV-19/India/MH-NCDC-3965/2020\|EPI_ISL_436444\|202 | .................... | .................... | ..................... | .................... | .................... | .................... |
| B | 3900 | hCoV-19/India/DL-NCDC-3982/2020\|EPI_ISL_436445\|202 | .................... | .................... | ..................... | .................... | .................... | .................... |
| B | 3901 | hCoV-19/India/KA-NCDC-4055/2020\|EPI_ISL_436447\|202 | .................... | .................... | ..................... | .................... | .................... | .................... |
| B | 3902 | hCoV-19/India/DL-NCDC-4138/2020\|EPI_ISL_436448\|202 | .................... | .................... | ..................... | .................... | .................... | .................... |
| B | 3903 | hCoV-19/India/BR-NCDC-4155/2020\|EPI_ISL_436449\|202 | .................... | .................... | ..................... | .................... | .................... | .................... |
| B | 3904 | hCoV-19/India/DL-NCDC-4189/2020\|EPI_ISL_436450\|202 | .................... | .................... | ..................... | .................... | .................... | .................... |
| B | 3905 | hCoV-19/India/DL-NCDC-4205/2020\|EPI_ISL_436451\|202 | .................... | .................... | ..................... | .................... | .................... | .................... |
| B | 3906 | hCoV-19/India/DL-NCDC-4208/2020\|EPI_ISL_436452\|202 | .................... | .................... | ..................... | .................... | .................... | .................... |
| B | 3907 | hCoV-19/India/MP-NCDC-4444/2020\|EPI_ISL_436453\|202 | .................... | .................... | ..................... | .................... | .................... | .................... |
| B | 3908 | hCoV-19/India/DL-NCDC-4475/2020\|EPI_ISL_436454\|202 | .................... | .................... | ..................... | .................... | .................... | .................... |
| B | 3909 | hCoV-19/India/DL-NCDC-4476/2020\|EPI_ISL_436455\|202 | .................... | .................... | ..................... | .................... | .................... | .................... |
| B | 3910 | hCoV-19/India/MP-NCDC-4846/2020\|EPI_ISL_436456\|202 | .................... | .................... | ..................... | .................... | .................... | .................... |
| B | 3911 | hCoV-19/India/MP-NCDC-4859/2020\|EPI_ISL_436457\|202 | .................... | .................... | ..................... | .................... | .................... | .................... |

| | | | | | | | | |
|---|---|---|---|---|---|---|---|---|
| B | 3912 | hCoV-19/India/MP-NCDC-4867/2020\|EPI_ISL_436458\|202 | ···················· | ···················· | ···················· | ···················· | ···················· | ···················· |
| B | 3913 | hCoV-19/India/MP-NCDC-4874/2020\|EPI_ISL_436459\|202 | ···················· | ···················· | ···················· | ···················· | ···················· | ···················· |
| B | 3914 | hCoV-19/India/MP-NCDC-4876/2020\|EPI_ISL_436460\|202 | ···················· | ···················· | ···················· | ···················· | ···················· | ···················· |
| B | 3915 | hCoV-19/India/MP-NCDC-4877/2020\|EPI_ISL_436461\|202 | ···················· | ···················· | ···················· | ···················· | ···················· | ···················· |
| B | 3916 | hCoV-19/India/MP-NCDC-4878/2020\|EPI_ISL_436462\|202 | ···················· | ···················· | ···················· | ···················· | ···················· | ···················· |
| B | 3917 | hCoV-19/India/MP-NCDC-4879/2020\|EPI_ISL_436463\|202 | ···················· | ···················· | ···················· | ···················· | ···················· | ···················· |
| B | 3918 | hCoV-19/USA/AK-PHL152/2020\|EPI_ISL_436464\|2020-04- | ···················· | ···················· | ···················· | ···················· | ···················· | ···················· |
| B | 3919 | hCoV-19/USA/PA-MGSC11-04/2020\|EPI_ISL_436466\|2020- | ···················· | ···················· | ···················· | ···················· | ···················· | ···················· |
| B | 3920 | hCoV-19/USA/PA-MGSC12-04/2020\|EPI_ISL_436467\|2020- | ···················· | ···················· | ···················· | ···················· | ···················· | ···················· |
| B | 3921 | hCoV-19/USA/PA-MGSC13-04/2020\|EPI_ISL_436468\|2020- | ···················· | ···················· | ···················· | ···················· | ···················· | ···················· |
| B | 3922 | hCoV-19/USA/PA-MGSC14-04/2020\|EPI_ISL_436469\|2020- | ···················· | ···················· | ···················· | ···················· | ···················· | ···················· |
| B | 3923 | hCoV-19/USA/PA-MGSC15-04/2020\|EPI_ISL_436470\|2020- | ···················· | ···················· | ···················· | ···················· | ···················· | ···················· |
| B | 3924 | hCoV-19/USA/PA-MGSC16-04/2020\|EPI_ISL_436471\|2020- | ···················· | ···················· | ···················· | ···················· | ···················· | ···················· |
| B | 3925 | hCoV-19/USA/PA-MGSC17-04/2020\|EPI_ISL_436472\|2020- | ···················· | ···················· | ···················· | ···················· | ···················· | ···················· |
| B | 3926 | hCoV-19/USA/PA-MGSC18-04/2020\|EPI_ISL_436473\|2020- | ···················· | ···················· | ···················· | ···················· | ···················· | ···················· |
| B | 3927 | hCoV-19/USA/PA-MGSC20-04/2020\|EPI_ISL_436474\|2020- | ···················· | ···················· | ···················· | ···················· | ···················· | ···················· |
| B | 3928 | hCoV-19/USA/PA-MGSC21-04/2020\|EPI_ISL_436475\|2020- | ···················· | ···················· | ···················· | ···················· | ···················· | ···················· |
| B | 3929 | hCoV-19/USA/PA-MGSC23-05/2020\|EPI_ISL_436476\|2020- | ···················· | ···················· | ···················· | ···················· | ···················· | ···················· |
| B | 3930 | hCoV-19/USA/PA-MGSC24-05/2020\|EPI_ISL_436477\|2020- | ···················· | ···················· | ···················· | ···················· | ···················· | ···················· |
| B | 3931 | hCoV-19/USA/PA-MGSC25-05/2020\|EPI_ISL_436478\|2020- | ···················· | ···················· | ···················· | ···················· | ···················· | ···················· |
| B | 3932 | hCoV-19/USA/PA-MGSC28-05/2020\|EPI_ISL_436479\|2020- | ···················· | ···················· | ···················· | ···················· | ···················· | ···················· |
| B | 3933 | hCoV-19/USA/PA-MGSC29-05/2020\|EPI_ISL_436480\|2020- | ···················· | ···················· | ···················· | ···················· | ···················· | ···················· |
| B | 3934 | hCoV-19/USA/PA-MGSC30-05/2020\|EPI_ISL_436481\|2020- | ···················· | ···················· | ···················· | ···················· | ···················· | ···················· |
| B | 3935 | hCoV-19/USA/PA-MGSC32-05/2020\|EPI_ISL_436482\|2020- | ···················· | ···················· | ···················· | ···················· | ···················· | ···················· |
| B | 3936 | hCoV-19/USA/PA-MGSC33-05/2020\|EPI_ISL_436483\|2020- | ···················· | ···················· | ···················· | ···················· | ···················· | ···················· |
| B | 3937 | hCoV-19/USA/PA-MGSC35-05/2020\|EPI_ISL_436484\|2020- | ···················· | ···················· | ···················· | ···················· | ···················· | ···················· |
| B | 3938 | hCoV-19/USA/PA-MGSC36-05/2020\|EPI_ISL_436485\|2020- | ···················· | ···················· | ···················· | ···················· | ···················· | ···················· |
| B | 3939 | hCoV-19/USA/PA-MGSC38-05/2020\|EPI_ISL_436486\|2020- | ···················· | ···················· | ···················· | ···················· | ···················· | ···················· |
| B | 3940 | hCoV-19/USA/PA-MGSC39-05/2020\|EPI_ISL_436487\|2020- | ···················· | ···················· | ···················· | ···················· | ···················· | ···················· |
| B | 3941 | hCoV-19/USA/PA-MGSC40-05/2020\|EPI_ISL_436488\|2020- | ···················· | ···················· | ···················· | ···················· | ···················· | ···················· |
| B | 3942 | hCoV-19/USA/PA-MGSC41-05/2020\|EPI_ISL_436489\|2020- | ···················· | ···················· | ···················· | ···················· | ···················· | ···················· |
| B | 3943 | hCoV-19/USA/PA-MGSC42-05/2020\|EPI_ISL_436490\|2020- | ···················· | ···················· | ···················· | ···················· | ···················· | ···················· |
| B | 3944 | hCoV-19/USA/PA-MGSC43-05/2020\|EPI_ISL_436491\|2020- | ···················· | ···················· | ···················· | ···················· | ···················· | ···················· |
| B | 3945 | hCoV-19/USA/PA-MGSC44-05/2020\|EPI_ISL_436492\|2020- | ···················· | ···················· | ···················· | ···················· | ···················· | ···················· |
| B | 3946 | hCoV-19/USA/PA-MGSC45-05/2020\|EPI_ISL_436493\|2020- | ···················· | ···················· | ···················· | ···················· | ···················· | ···················· |
| B | 3947 | hCoV-19/USA/PA-MGSC46-05/2020\|EPI_ISL_436494\|2020- | ···················· | ···················· | ···················· | ···················· | ···················· | ···················· |
| B | 3948 | hCoV-19/USA/PA-MGSC47-05/2020\|EPI_ISL_436495\|2020- | ···················· | ···················· | ···················· | ···················· | ···················· | ···················· |
| B | 3949 | hCoV-19/USA/PA-MGSC48-05/2020\|EPI_ISL_436496\|2020- | ···················· | ···················· | ···················· | ···················· | ···················· | ···················· |
| B | 3950 | hCoV-19/USA/PA-MGSC49-05/2020\|EPI_ISL_436497\|2020- | ···················· | ···················· | ···················· | ···················· | ···················· | ···················· |
| B | 3951 | hCoV-19/USA/PA-MGSC50-05/2020\|EPI_ISL_436498\|2020- | ···················· | ···················· | ···················· | ···················· | ···················· | ···················· |
| B | 3952 | hCoV-19/USA/PA-MGSC51-05/2020\|EPI_ISL_436499\|2020- | ···················· | ···················· | ···················· | ···················· | ···················· | ···················· |
| B | 3953 | hCoV-19/Saudi Arabia/KAUST-Makkah177/2020\|EPI_ISL_ | ···················· | ···················· | ···················· | ···················· | ···················· | ···················· |
| B | 3954 | hCoV-19/Saudi Arabia/KAUST-Makkah178/2020\|EPI_ISL_ | ···················· | ···················· | ···················· | ···················· | ···················· | ···················· |
| B | 3955 | hCoV-19/Saudi Arabia/KAUST-Makkah179/2020\|EPI_ISL_ | ···················· | ···················· | ···················· | ···················· | ···················· | ···················· |
| B | 3956 | hCoV-19/Saudi Arabia/KAUST-Makkah181/2020\|EPI_ISL_ | ···················· | ···················· | ···················· | ···················· | ···················· | ···················· |
| B | 3957 | hCoV-19/Saudi Arabia/KAUST-Makkah183/2020\|EPI_ISL_ | ···················· | ···················· | ···················· | ···················· | ···················· | ···················· |
| B | 3958 | hCoV-19/Saudi Arabia/KAUST-Makkah188/2020\|EPI_ISL_ | ···················· | ···················· | ···················· | ···················· | ···················· | ···················· |
| B | 3959 | hCoV-19/Saudi Arabia/KAUST-Makkah189/2020\|EPI_ISL_ | ···················· | ·A·················· | ···················· | ···················· | ···················· | ··A················· |
| B | 3960 | hCoV-19/Saudi Arabia/KAUST-Makkah190/2020\|EPI_ISL_ | ···················· | ···················· | ···················· | ···················· | ···················· | ···················· |
| B | 3961 | hCoV-19/Saudi Arabia/KAUST-Makkah193/2020\|EPI_ISL_ | ···················· | ···················· | ···················· | ···················· | ···················· | ···················· |
| B | 3962 | hCoV-19/Saudi Arabia/KAUST-Makkah194/2020\|EPI_ISL_ | ···················· | ···················· | ···················· | ···················· | ···················· | ···················· |
| B | 3963 | hCoV-19/Saudi Arabia/KAUST-Makkah196/2020\|EPI_ISL_ | ···················· | ·A·················· | ···················· | ···················· | ···················· | ··A················· |
| B | 3964 | hCoV-19/Saudi Arabia/KAUST-Makkah198/2020\|EPI_ISL_ | ···················· | ···················· | ···················· | ···················· | ···················· | ···················· |
| B | 3965 | hCoV-19/Saudi Arabia/KAUST-Makkah202/2020\|EPI_ISL_ | ···················· | ···················· | ···················· | ···················· | ···················· | ···················· |
| B | 3966 | hCoV-19/Saudi Arabia/KAUST-Makkah203/2020\|EPI_ISL_ | ···················· | ···················· | ···················· | ···················· | ···················· | ···················· |
| B | 3967 | hCoV-19/Saudi Arabia/KAUST-Makkah204/2020\|EPI_ISL_ | ···················· | ···················· | ···················· | ···················· | ···················· | ···················· |
| B | 3968 | hCoV-19/Saudi Arabia/KAUST-Madinah208/2020\|EPI_ISL | ···················· | ·A·················· | ···················· | ···················· | ···················· | ··A················· |
| B | 3969 | hCoV-19/Saudi Arabia/KAUST-Madinah211/2020\|EPI_ISL | ···················· | ···················· | ···················· | ···················· | ···················· | ···················· |
| B | 3970 | hCoV-19/Saudi Arabia/KAUST-Madinah213/2020\|EPI_ISL | ···················· | ···················· | ···················· | ···················· | ···················· | ···················· |
| B | 3971 | hCoV-19/Saudi Arabia/KAUST-Makkah215/2020\|EPI_ISL_ | ···················· | ···················· | ···················· | ···················· | ···················· | ···················· |
| B | 3972 | hCoV-19/Saudi Arabia/KAUST-Makkah217/2020\|EPI_ISL_ | ···················· | ···················· | ···················· | ···················· | ···················· | ···················· |
| B | 3973 | hCoV-19/Saudi Arabia/KAUST-Makkah218/2020\|EPI_ISL_ | ···················· | ···················· | ···················· | ···················· | ···················· | ···················· |
| B | 3974 | hCoV-19/Saudi Arabia/KAUST-Makkah220/2020\|EPI_ISL_ | ···················· | ···················· | ···················· | ···················· | ···················· | ···················· |
| B | 3975 | hCoV-19/Saudi Arabia/KAUST-Makkah225/2020\|EPI_ISL_ | ···················· | ···················· | ···················· | ···················· | ···················· | ···················· |
| B | 3976 | hCoV-19/Saudi Arabia/KAUST-Makkah227/2020\|EPI_ISL_ | ···················· | ·A·················· | ···················· | ···················· | ···················· | ··A················· |
| B | 3977 | hCoV-19/Saudi Arabia/KAUST-Makkah228/2020\|EPI_ISL_ | ···················· | ···················· | ···················· | ···················· | ···················· | ···················· |
| B | 3978 | hCoV-19/Saudi Arabia/KAUST-Madinah264/2020\|EPI_ISL | ···················· | ···················· | ···················· | ···················· | ···················· | ···················· |
| B | 3979 | hCoV-19/Saudi Arabia/KAUST-Madinah265/2020\|EPI_ISL | ···················· | ···················· | ···················· | ···················· | ···················· | ··A················· |
| B | 3980 | hCoV-19/Saudi Arabia/KAUST-Madinah266/2020\|EPI_ISL | ···················· | ···················· | ···················· | ···················· | ···················· | ···················· |
| B | 3981 | hCoV-19/Saudi Arabia/KAUST-Jeddah68/2020\|EPI_ISL_4 | ···················· | ···················· | ···················· | ···················· | ···················· | ···················· |
| B | 3982 | hCoV-19/USA/VA-DCLS-0130/2020\|EPI_ISL_437769\|2020- | ···················· | ···················· | ···················· | ···················· | ···················· | ···················· |
| B | 3983 | hCoV-19/USA/VA-DCLS-0131/2020\|EPI_ISL_437770\|2020- | ···················· | ···················· | ···················· | ···················· | ···················· | ···················· |
| B | 3984 | hCoV-19/USA/VA-DCLS-0132/2020\|EPI_ISL_437771\|2020- | ···················· | ···················· | ···················· | ···················· | ···················· | ···················· |
| B | 3985 | hCoV-19/USA/VA-DCLS-0133/2020\|EPI_ISL_437772\|2020- | ···················· | ···················· | ···················· | ···················· | ···················· | ···················· |
| B | 3986 | hCoV-19/USA/VA-DCLS-0134/2020\|EPI_ISL_437773\|2020- | ···················· | ···················· | ···················· | ···················· | ···················· | ···················· |
| B | 3987 | hCoV-19/USA/VA-DCLS-0135/2020\|EPI_ISL_437774\|2020- | ···················· | ···················· | ···················· | ···················· | ···················· | ···················· |
| B | 3988 | hCoV-19/USA/VA-DCLS-0136/2020\|EPI_ISL_437775\|2020- | ···················· | ···················· | ···················· | ···················· | ···················· | ···················· |
| B | 3989 | hCoV-19/USA/VA-DCLS-0137/2020\|EPI_ISL_437776\|2020- | ···················· | ···················· | ···················· | ···················· | ···················· | ···················· |

| | | | | | | | |
|---|---|---|---|---|---|---|---|
| B | 3990 hCoV-19/USA/VA-DCLS-0138/2020\|EPI_ISL_437777\|2020- | ···················· | ···················· | ····················· | ···················· | ···················· | ···················· |
| B | 3991 hCoV-19/USA/VA-DCLS-0139/2020\|EPI_ISL_437778\|2020- | ···················· | ···················· | ····················· | ···················· | ···················· | ···················· |
| B | 3992 hCoV-19/USA/VA-DCLS-0140/2020\|EPI_ISL_437779\|2020- | ···················· | ···················· | ····················· | ···················· | ···················· | ···················· |
| B | 3993 hCoV-19/USA/VA-DCLS-0141/2020\|EPI_ISL_437780\|2020- | ···················· | ···················· | ····················· | ···················· | ···················· | ···················· |
| B | 3994 hCoV-19/USA/VA-DCLS-0142/2020\|EPI_ISL_437781\|2020- | ···················· | ···················· | ····················· | ···················· | ···················· | ···················· |
| B | 3995 hCoV-19/USA/VA-DCLS-0144/2020\|EPI_ISL_437782\|2020- | ···················· | ···················· | ····················· | ···················· | ···················· | ···················· |
| B | 3996 hCoV-19/USA/VA-DCLS-0146/2020\|EPI_ISL_437783\|2020- | ···················· | ···················· | ····················· | ···················· | ···················· | ···················· |
| B | 3997 hCoV-19/USA/VA-DCLS-0151/2020\|EPI_ISL_437784\|2020- | ···················· | ···················· | ····················· | ···················· | ···················· | ···················· |
| B | 3998 hCoV-19/USA/VA-DCLS-0152/2020\|EPI_ISL_437785\|2020- | ···················· | ···················· | ····················· | ···················· | ···················· | ···················· |
| B | 3999 hCoV-19/USA/VA-DCLS-0153/2020\|EPI_ISL_437786\|2020- | ···················· | ···················· | ····················· | ···················· | ···················· | ···················· |
| B | 4000 hCoV-19/USA/VA-DCLS-0155/2020\|EPI_ISL_437788\|2020- | ···················· | ···················· | ····················· | ···················· | ···················· | ···················· |
| B | 4001 hCoV-19/USA/VA-DCLS-0156/2020\|EPI_ISL_437789\|2020- | ···················· | ···················· | ····················· | ···················· | ···················· | ···················· |
| B | 4002 hCoV-19/USA/VA-DCLS-0157/2020\|EPI_ISL_437790\|2020- | ···················· | ···················· | ····················· | ···················· | ···················· | ···················· |
| B | 4003 hCoV-19/USA/VA-DCLS-0158/2020\|EPI_ISL_437791\|2020- | ···················· | ···················· | ····················· | ···················· | ···················· | ···················· |
| B | 4004 hCoV-19/USA/VA-DCLS-0159/2020\|EPI_ISL_437792\|2020- | ···················· | ···················· | ····················· | ···················· | ···················· | ···················· |
| B | 4005 hCoV-19/USA/VA-DCLS-0160/2020\|EPI_ISL_437793\|2020- | ···················· | ···················· | ····················· | ···················· | ···················· | ···················· |
| B | 4006 hCoV-19/USA/VA-DCLS-0161/2020\|EPI_ISL_437794\|2020- | ···················· | ···················· | ····················· | ···················· | ···················· | ···················· |
| B | 4007 hCoV-19/USA/VA-DCLS-0162/2020\|EPI_ISL_437795\|2020- | ···················· | ···················· | ····················· | ···················· | ···················· | ···················· |
| B | 4008 hCoV-19/USA/VA-DCLS-0163/2020\|EPI_ISL_437796\|2020- | ···················· | ···················· | ····················· | ···················· | ···················· | ···················· |
| B | 4009 hCoV-19/USA/VA-DCLS-0164/2020\|EPI_ISL_437797\|2020- | ···················· | ···················· | ····················· | ···················· | ···················· | ···················· |
| B | 4010 hCoV-19/USA/VA-DCLS-0165/2020\|EPI_ISL_437798\|2020- | ···················· | ···················· | ····················· | ···················· | ···················· | ···················· |
| B | 4011 hCoV-19/USA/VA-DCLS-0166/2020\|EPI_ISL_437799\|2020- | ···················· | ···················· | ····················· | ···················· | ···················· | ···················· |
| B | 4012 hCoV-19/England/CAMB-7C6D0/2020\|EPI_ISL_442281\|202 | ···················· | ···················· | ····················· | ···················· | ···················· | ···················· |
| B | 4013 hCoV-19/England/CAMB-7C791/2020\|EPI_ISL_442282\|202 | ···················· | ···················· | ····················· | ···················· | ···················· | ···················· |
| B | 4014 hCoV-19/England/CAMB-7B783/2020\|EPI_ISL_442283\|202 | ···················· | ···················· | ····················· | ···················· | ···················· | ···················· |
| B | 4015 hCoV-19/England/CAMB-7B756/2020\|EPI_ISL_442284\|202 | ···················· | ···················· | ····················· | ···················· | ···················· | ···················· |
| B | 4016 hCoV-19/England/CAMB-7D332/2020\|EPI_ISL_442285\|202 | ···················· | ···················· | ····················· | ···················· | ···················· | ···················· |
| B | 4017 hCoV-19/England/CAMB-7C5E2/2020\|EPI_ISL_442286\|202 | ···················· | ···················· | ····················· | ···················· | ···················· | ···················· |
| B | 4018 hCoV-19/England/CAMB-7B880/2020\|EPI_ISL_442287\|202 | ···················· | ·············A······ | ····················· | ···················· | ···················· | ···················· |
| B | 4019 hCoV-19/England/CAMB-7C61C/2020\|EPI_ISL_442288\|202 | ···················· | ···················· | ····················· | ···················· | ···················· | ···················· |
| B | 4020 hCoV-19/England/CAMB-7D147/2020\|EPI_ISL_442289\|202 | ···················· | ···················· | ····················· | ···················· | ···················· | ···················· |
| B | 4021 hCoV-19/England/CAMB-7BB4B/2020\|EPI_ISL_442291\|202 | ···················· | ···················· | ····················· | ···················· | ···················· | ···················· |
| B | 4022 hCoV-19/England/CAMB-7B8EA/2020\|EPI_ISL_442292\|202 | ···················· | ···················· | ····················· | ···················· | ···················· | ···················· |
| B | 4023 hCoV-19/England/CAMB-7DFA7/2020\|EPI_ISL_442294\|202 | ···················· | ···················· | ····················· | ···················· | ···················· | ···················· |
| B | 4024 hCoV-19/England/CAMB-7BA3F/2020\|EPI_ISL_442295\|202 | ···················· | ···················· | ····················· | ···················· | ···················· | ···················· |
| B | 4025 hCoV-19/England/CAMB-7D3BA/2020\|EPI_ISL_442296\|202 | ···················· | ···················· | ····················· | ···················· | ···················· | ···················· |
| B | 4026 hCoV-19/England/CAMB-7D736/2020\|EPI_ISL_442297\|202 | ···················· | ···················· | ····················· | ···················· | ···················· | ···················· |
| B | 4027 hCoV-19/England/CAMB-7CD62/2020\|EPI_ISL_442299\|202 | ···················· | ···················· | ····················· | ···················· | ···················· | ···················· |
| B | 4028 hCoV-19/England/CAMB-82E0C/2020\|EPI_ISL_443600\|202 | ···················· | ···················· | ····················· | ···················· | ···················· | ···················· |
| B | 4029 hCoV-19/England/CAMB-84408/2020\|EPI_ISL_443602\|202 | ···················· | ···················· | ····················· | ···················· | ···················· | ···················· |
| B | 4030 hCoV-19/England/CAMB-1AA553/2020\|EPI_ISL_443604\|20 | ···················· | ···················· | ····················· | ···················· | ···················· | ···················· |
| B | 4031 hCoV-19/England/CAMB-8308D/2020\|EPI_ISL_443605\|202 | ···················· | ···················· | ····················· | ···················· | ···················· | ···················· |
| B | 4032 hCoV-19/England/CAMB-83700/2020\|EPI_ISL_443607\|202 | ···················· | ···················· | ····················· | ···················· | ···················· | ···················· |
| B | 4033 hCoV-19/England/CAMB-834FA/2020\|EPI_ISL_443608\|202 | ···················· | ···················· | ····················· | ···················· | ···················· | ···················· |
| B | 4034 hCoV-19/England/CAMB-825BC/2020\|EPI_ISL_443609\|202 | ···················· | ···················· | ····················· | ···················· | ···················· | ···················· |
| B | 4035 hCoV-19/England/CAMB-83014/2020\|EPI_ISL_443612\|202 | ···················· | ···················· | ····················· | ···················· | ···················· | ···················· |
| B | 4036 hCoV-19/England/CAMB-82E39/2020\|EPI_ISL_443613\|202 | ···················· | ···················· | ····················· | ···················· | ···················· | ···················· |
| B | 4037 hCoV-19/England/CAMB-84356/2020\|EPI_ISL_443614\|202 | ···················· | ···················· | ····················· | ···················· | ···················· | ···················· |
| B | 4038 hCoV-19/England/CAMB-845BA/2020\|EPI_ISL_443616\|202 | ···················· | ·A·················· | ····················· | ···················· | ···················· | ···················· |
| B | 4039 hCoV-19/England/CAMB-8365E/2020\|EPI_ISL_443617\|202 | ···················· | ·A·················· | ····················· | ···················· | ···················· | ···················· |
| B | 4040 hCoV-19/England/CAMB-832B4/2020\|EPI_ISL_443618\|202 | ···················· | ···················· | ····················· | ···················· | ···················· | ···················· |
| B | 4041 hCoV-19/Denmark/ALAB-SSI-835/2020\|EPI_ISL_444930\|2 | ···················· | ···················· | ····················· | ···················· | ···················· | ···················· |
| B | 4042 hCoV-19/Denmark/ALAB-SSI-837/2020\|EPI_ISL_444931\|2 | ···················· | ···················· | ····················· | ···················· | ···················· | ···················· |
| B | 4043 hCoV-19/Denmark/ALAB-SSI-840/2020\|EPI_ISL_444933\|2 | ···················· | ···················· | ····················· | ···················· | ···················· | ···················· |
| B | 4044 hCoV-19/Denmark/ALAB-SSI-842/2020\|EPI_ISL_444934\|2 | ···················· | ···················· | ····················· | ···················· | ···················· | ···················· |
| B | 4045 hCoV-19/Denmark/ALAB-SSI-843/2020\|EPI_ISL_444935\|2 | ···················· | ···················· | ····················· | ···················· | ···················· | ···················· |
| B | 4046 hCoV-19/Denmark/ALAB-SSI-846/2020\|EPI_ISL_444937\|2 | ···················· | ···················· | ····················· | ···················· | ···················· | ···················· |
| B | 4047 hCoV-19/Denmark/ALAB-SSI-847/2020\|EPI_ISL_444938\|2 | ···················· | ···················· | ····················· | ···················· | ···················· | ···················· |
| B | 4048 hCoV-19/Denmark/ALAB-SSI-848/2020\|EPI_ISL_444939\|2 | ···················· | ···················· | ····················· | ···················· | ···················· | ···················· |
| B | 4049 hCoV-19/Denmark/ALAB-SSI-852/2020\|EPI_ISL_444941\|2 | ···················· | ···················· | ····················· | ···················· | ···················· | ···················· |
| B | 4050 hCoV-19/Denmark/ALAB-SSI-854/2020\|EPI_ISL_444942\|2 | ···················· | ···················· | ····················· | ···················· | ···················· | ···················· |
| B | 4051 hCoV-19/Denmark/ALAB-SSI-856/2020\|EPI_ISL_444944\|2 | ···················· | ···················· | ····················· | ···················· | ···················· | ···················· |
| B | 4052 hCoV-19/Denmark/ALAB-SSI-860/2020\|EPI_ISL_444945\|2 | ···················· | ···················· | ····················· | ···················· | ···················· | ···················· |
| B | 4053 hCoV-19/Denmark/ALAB-SSI-861/2020\|EPI_ISL_444946\|2 | ···················· | ···················· | ····················· | ···················· | ···················· | ···················· |
| B | 4054 hCoV-19/Denmark/ALAB-SSI-862/2020\|EPI_ISL_444947\|2 | ···················· | ···················· | ····················· | ···················· | ···················· | ···················· |
| B | 4055 hCoV-19/Denmark/ALAB-SSI-866/2020\|EPI_ISL_444948\|2 | ···················· | ···················· | ····················· | ···················· | ···················· | ···················· |
| B | 4056 hCoV-19/Denmark/ALAB-SSI-867/2020\|EPI_ISL_444949\|2 | ···················· | ···················· | ····················· | ···················· | ···················· | ···················· |
| B | 4057 hCoV-19/Denmark/ALAB-SSI-868/2020\|EPI_ISL_444950\|2 | ···················· | ···················· | ····················· | ···················· | ···················· | ···················· |
| B | 4058 hCoV-19/USA/MI-MDHHS-SC20304/2020\|EPI_ISL_447101\|2 | ···················· | ···················· | ····················· | ···················· | ···················· | ···················· |
| B | 4059 hCoV-19/USA/MI-MDHHS-SC20305/2020\|EPI_ISL_447102\|2 | ···················· | ···················· | ····················· | ···················· | ···················· | ···················· |
| B | 4060 hCoV-19/USA/MI-MDHHS-SC20307/2020\|EPI_ISL_447104\|2 | ···················· | ···················· | ····················· | ···················· | ···················· | ···················· |
| B | 4061 hCoV-19/USA/MI-MDHHS-SC20308/2020\|EPI_ISL_447105\|2 | ···················· | ···················· | ····················· | ···················· | ···················· | ···················· |
| B | 4062 hCoV-19/USA/MI-MDHHS-SC20309/2020\|EPI_ISL_447106\|2 | ···················· | ···················· | ····················· | ···················· | ···················· | ···················· |
| B | 4063 hCoV-19/USA/MI-MDHHS-SC20310/2020\|EPI_ISL_447107\|2 | ···················· | ···················· | ····················· | ···················· | ···················· | ···················· |
| B | 4064 hCoV-19/USA/MI-MDHHS-SC20314/2020\|EPI_ISL_447111\|2 | ···················· | ···················· | ····················· | ···················· | ···················· | ···················· |
| B | 4065 hCoV-19/USA/MI-MDHHS-SC20316/2020\|EPI_ISL_447113\|2 | ···················· | ···················· | ····················· | ···················· | ···················· | ···················· |
| B | 4066 hCoV-19/USA/MI-MDHHS-SC20318/2020\|EPI_ISL_447115\|2 | ···················· | ···················· | ····················· | ···················· | ···················· | ···················· |
| B | 4067 hCoV-19/USA/MI-MDHHS-SC20319/2020\|EPI_ISL_447116\|2 | ···················· | ···················· | ····················· | ···················· | ···················· | ···················· |

| | | | | | | | | |
|---|---|---|---|---|---|---|---|---|
| B | 4068 | hCoV-19/USA/MI-MDHHS-SC20320/2020\|EPI_ISL_447117\|2 | ···················· | ···················· | ····················· | ···················· | ···················· | ···················· |
| B | 4069 | hCoV-19/USA/MI-MDHHS-SC20321/2020\|EPI_ISL_447118\|2 | ···················· | ···················· | ····················· | ···················· | ···················· | ···················· |
| B | 4070 | hCoV-19/Chile/AR-ISPCH-9/2020\|EPI_ISL_447119\|2020- | ···················· | ···················· | ····················· | ···················· | ···················· | ···················· |
| B | 4071 | hCoV-19/Belgium/ULG-6839/2020\|EPI_ISL_447120\|2020- | ···················· | ···················· | ····················· | ···················· | ···················· | ···················· |
| B | 4072 | hCoV-19/Belgium/ULG-6842/2020\|EPI_ISL_447121\|2020- | ···················· | ···················· | ····················· | ···················· | ···················· | ···················· |
| B | 4073 | hCoV-19/Belgium/ULG-8937/2020\|EPI_ISL_447122\|2020- | ···················· | ···················· | ····················· | ···················· | ···················· | ···················· |
| B | 4074 | hCoV-19/Belgium/ULG-9535/2020\|EPI_ISL_447123\|2020- | ···················· | ···················· | ····················· | ···················· | ···················· | ···················· |
| B | 4075 | hCoV-19/Belgium/ULG-9536/2020\|EPI_ISL_447124\|2020- | ···················· | ···················· | ····················· | ···················· | ···················· | ···················· |
| B | 4076 | hCoV-19/Belgium/ULG-0184/2020\|EPI_ISL_447127\|2020- | ···················· | ···················· | ····················· | ···················· | ···················· | ···················· |
| B | 4077 | hCoV-19/Belgium/ULG-0186/2020\|EPI_ISL_447128\|2020- | ···················· | ···················· | ····················· | ···················· | ···················· | ···················· |
| B | 4078 | hCoV-19/Belgium/ULG-0189/2020\|EPI_ISL_447129\|2020- | ···················· | ···················· | ····················· | ···················· | ···················· | ···················· |
| B | 4079 | hCoV-19/Belgium/ULG-0190/2020\|EPI_ISL_447130\|2020- | ···················· | ···················· | ····················· | ···················· | ···················· | ···················· |
| B | 4080 | hCoV-19/Belgium/ULG-0536/2020\|EPI_ISL_447133\|2020- | ···················· | ···················· | ····················· | ···················· | ···················· | ···················· |
| B | 4081 | hCoV-19/Belgium/ULG-0544/2020\|EPI_ISL_447134\|2020- | ···················· | ···················· | ····················· | ···················· | ···················· | ···················· |
| B | 4082 | hCoV-19/Belgium/ULG-0546/2020\|EPI_ISL_447135\|2020- | ···················· | ···················· | ····················· | ···················· | ···················· | ···················· |
| B | 4083 | hCoV-19/Belgium/ULG-7921/2020\|EPI_ISL_447136\|2020- | ···················· | ···················· | ····················· | ···················· | ···················· | ···················· |
| B | 4084 | hCoV-19/Belgium/ULG-10145/2020\|EPI_ISL_447137\|2020 | ···················· | ···················· | ····················· | ···················· | ···················· | ···················· |
| B | 4085 | hCoV-19/Belgium/ULG-10146/2020\|EPI_ISL_447138\|2020 | ···················· | ···················· | ····················· | ···················· | ···················· | ···················· |
| B | 4086 | hCoV-19/Belgium/ULG-10169/2020\|EPI_ISL_447139\|2020 | ···················· | ···················· | ····················· | ···················· | ···················· | ···················· |
| B | 4087 | hCoV-19/Belgium/ULG-10152/2020\|EPI_ISL_447140\|2020 | ···················· | ···················· | ····················· | ···················· | ···················· | ···················· |
| B | 4088 | hCoV-19/Belgium/ULG-0183/2020\|EPI_ISL_447141\|2020- | ···················· | ···················· | ····················· | ···················· | ···················· | ···················· |
| B | 4089 | hCoV-19/Belgium/ULG-10151/2020\|EPI_ISL_447148\|2020 | ···················· | ···················· | ····················· | ···················· | ···················· | ···················· |
| B | 4090 | hCoV-19/Belgium/ULG-10156/2020\|EPI_ISL_447152\|2020 | ···················· | ···················· | ····················· | ···················· | ···················· | ···················· |
| B | 4091 | hCoV-19/Belgium/ULG-10158/2020\|EPI_ISL_447153\|2020 | ···················· | ···················· | ····················· | ···················· | ···················· | ···················· |
| B | 4092 | hCoV-19/Belgium/ULG-10160/2020\|EPI_ISL_447155\|2020 | ···················· | ···················· | ····················· | ···················· | ···················· | ···················· |
| B | 4093 | hCoV-19/USA/MI-MDHHS-SC20191/2020\|EPI_ISL_447163\|2 | ···················· | ···················· | ····················· | ···················· | ···················· | ···················· |
| B | 4094 | hCoV-19/USA/MI-MDHHS-SC20194/2020\|EPI_ISL_447165\|2 | ···················· | ···················· | ····················· | ···················· | ···················· | ···················· |
| B | 4095 | hCoV-19/USA/MI-MDHHS-SC20196/2020\|EPI_ISL_447167\|2 | ···················· | ···················· | ····················· | ···················· | ···················· | ···················· |
| B | 4096 | hCoV-19/USA/MI-MDHHS-SC20197/2020\|EPI_ISL_447168\|2 | ···················· | ···················· | ····················· | ···················· | ···················· | ···················· |
| B | 4097 | hCoV-19/USA/NY-NYUMC373/2020\|EPI_ISL_430400\|2020-0 | ···················· | ···················· | ····················· | ···················· | ···················· | ···················· |
| B | 4098 | hCoV-19/USA/CT-NYUMC374/2020\|EPI_ISL_430401\|2020-0 | ···················· | ···················· | ····················· | ···················· | ···················· | ···················· |
| B | 4099 | hCoV-19/USA/NY-NYUMC375/2020\|EPI_ISL_430402\|2020-0 | ···················· | ···················· | ····················· | ···················· | ···················· | ···················· |
| B | 4100 | hCoV-19/USA/NJ-NYUMC376/2020\|EPI_ISL_430403\|2020-0 | ···················· | ···················· | ····················· | ···················· | ···················· | ···················· |
| B | 4101 | hCoV-19/USA/NY-NYUMC377/2020\|EPI_ISL_430404\|2020-0 | ···················· | ···················· | ····················· | ···················· | ···················· | ···················· |
| B | 4102 | hCoV-19/USA/NJ-NYUMC378/2020\|EPI_ISL_430405\|2020-0 | ···················· | ···················· | ····················· | ···················· | ···················· | ···················· |
| B | 4103 | hCoV-19/USA/NY-NYUMC379/2020\|EPI_ISL_430406\|2020-0 | ···················· | ···················· | ····················· | ···················· | ···················· | ···················· |
| B | 4104 | hCoV-19/USA/NY-NYUMC380/2020\|EPI_ISL_430407\|2020-0 | ···················· | ···················· | ····················· | ···················· | ···················· | ···················· |
| B | 4105 | hCoV-19/USA/NY-NYUMC381/2020\|EPI_ISL_430408\|2020-0 | ···················· | ···················· | ····················· | ···················· | ···················· | ···················· |
| B | 4106 | hCoV-19/USA/NY-NYUMC382/2020\|EPI_ISL_430409\|2020-0 | ···················· | ···················· | ····················· | ···················· | ···················· | ···················· |
| B | 4107 | hCoV-19/USA/NY-NYUMC405/2020\|EPI_ISL_430432\|2020-0 | ···················· | ···················· | ····················· | ···················· | ···················· | ···················· |
| B | 4108 | hCoV-19/USA/NY-NYUMC406/2020\|EPI_ISL_430433\|2020-0 | ···················· | ···················· | ····················· | ···················· | ···················· | ···················· |
| B | 4109 | hCoV-19/USA/NY-NYUMC407/2020\|EPI_ISL_430434\|2020-0 | ···················· | ···················· | ····················· | ···················· | ···················· | ···················· |
| B | 4110 | hCoV-19/Malaysia/IMR_WC1097/2020\|EPI_ISL_430441\|20 | ···················· | ···················· | ····················· | ···················· | ···················· | ···················· |
| B | 4111 | hCoV-19/Malaysia/IMR_WC1098/2020\|EPI_ISL_430442\|20 | ···················· | ···················· | ····················· | ···················· | ···················· | ···················· |
| B | 4112 | hCoV-19/India/WB-S3/2020\|EPI_ISL_430464\|2020-03-21 | ···················· | ···················· | ····················· | ···················· | ···················· | ···················· |
| B | 4113 | hCoV-19/India/WB-S5/2020\|EPI_ISL_430465\|2020-03-28 | ···················· | ···················· | ····················· | ···················· | ···················· | ···················· |
| B | 4114 | hCoV-19/India/WB-S6/2020\|EPI_ISL_430466\|2020-03-26 | ···················· | ···················· | ····················· | ···················· | ···················· | ···················· |
| B | 4115 | hCoV-19/India/WB-S11/2020\|EPI_ISL_430467\|2020-04-0 | ···················· | ···················· | ····················· | ···················· | ···················· | ···················· |
| B | 4116 | hCoV-19/India/WB-S2/2020\|EPI_ISL_430468\|2020-03-21 | ···················· | ···················· | ····················· | ···················· | ···················· | ···················· |
| B | 4117 | hCoV-19/Greece/127_HPI/2020\|EPI_ISL_430469\|2020-02 | ···················· | ···················· | ····················· | ···················· | ···················· | ···················· |
| B | 4132 | hCoV-19/Fujian/3520022Y/2020\|EPI_ISL_431785\|2020-0 | ···················· | ···················· | ····················· | ···················· | ···················· | ···················· |
| B | 4133 | hCoV-19/India/UP-NCDC-02321/2020\|EPI_ISL_435100\|20 | ···················· | ···················· | ····················· | ···················· | ···················· | ···················· |
| B | 4134 | hCoV-19/India/LA-NCDC-01441/2020\|EPI_ISL_435101\|20 | ···················· | ···················· | ····················· | ···················· | ···················· | ···················· |
| B | 4135 | hCoV-19/India/LA-NCDC-01444/2020\|EPI_ISL_435102\|20 | ···················· | ···················· | ····················· | ···················· | ···················· | ···················· |
| B | 4136 | hCoV-19/India/LA-NCDC-01604/2020\|EPI_ISL_435103\|20 | ···················· | ···················· | ····················· | ···················· | ···········N········ | ···················· |
| B | 4137 | hCoV-19/India/LA-NCDC-01614/2020\|EPI_ISL_435104\|20 | ···················· | ···················· | ····················· | ···················· | ···················· | ···················· |
| B | 4138 | hCoV-19/India/LA-NCDC-01616/2020\|EPI_ISL_435105\|20 | ···················· | ···················· | ····················· | ···················· | ···················· | ···················· |
| B | 4139 | hCoV-19/India/LA-NCDC-01760/2020\|EPI_ISL_435106\|20 | ···················· | ···················· | ····················· | ···················· | ···················· | ···················· |
| B | 4140 | hCoV-19/Spain/VC-FISABIO-316/2020\|EPI_ISL_436411\|2 | ···················· | ···················· | ····················· | ···················· | ···················· | ···················· |
| B | 4141 | hCoV-19/DRC/2299/2020\|EPI_ISL_436412\|2020-04-14 | ···················· | ···················· | ····················· | ···················· | ···················· | ···················· |
| B | 4142 | hCoV-19/India/UP-NCDC-2443/2020\|EPI_ISL_436413\|202 | ···················· | ···················· | ····················· | ···················· | ···················· | ···················· |
| B | 4143 | hCoV-19/India/WB-NCDC-2502/2020\|EPI_ISL_436414\|202 | ···················· | ···················· | ····················· | ···················· | ···················· | ···················· |
| B | 4144 | hCoV-19/India/DL-NCDC-2507/2020\|EPI_ISL_436415\|202 | ···················· | ···················· | ····················· | ···················· | ···················· | ···················· |
| B | 4145 | hCoV-19/India/RJ-NCDC-2522/2020\|EPI_ISL_436420\|202 | ···················· | ···················· | ····················· | ···················· | ···················· | ···················· |
| B | 4146 | hCoV-19/India/AS-NCDC-2525/2020\|EPI_ISL_436421\|202 | ···················· | ···················· | ····················· | ···················· | ···················· | ···················· |
| B | 4147 | hCoV-19/India/AS-NCDC-2535/2020\|EPI_ISL_436422\|202 | ···················· | ···················· | ····················· | ···················· | ···················· | ···················· |
| B | 4148 | hCoV-19/India/DL-NCDC-3163/2020\|EPI_ISL_436424\|202 | ···················· | ···················· | ····················· | ···················· | ···················· | ···················· |
| B | 4149 | hCoV-19/India/DL-NCDC-3175/2020\|EPI_ISL_436426\|202 | ···················· | ···················· | ····················· | ···················· | ···················· | ···················· |
| B | 4150 | hCoV-19/India/DL-NCDC-3181/2020\|EPI_ISL_436428\|202 | ···················· | ···················· | ····················· | ···················· | ···················· | ···················· |
| B | 4151 | hCoV-19/India/DL-NCDC-3190/2020\|EPI_ISL_436429\|202 | ···················· | ···················· | ····················· | ···················· | ···················· | ···················· |
| B | 4152 | hCoV-19/India/DL-NCDC-3195/2020\|EPI_ISL_436430\|202 | ···················· | ···················· | ····················· | ···················· | ···················· | ···················· |
| B | 4153 | hCoV-19/India/DL-NCDC-3230/2020\|EPI_ISL_436431\|202 | ···················· | ···················· | ····················· | ···················· | ···················· | ···················· |
| B | 4154 | hCoV-19/India/DL-NCDC-3264/2020\|EPI_ISL_436432\|202 | ···················· | ···················· | ····················· | ···················· | ·······N···W········ | ···················· |
| B | 4155 | hCoV-19/India/DL-NCDC-3271/2020\|EPI_ISL_436433\|202 | ···················· | ···················· | ····················· | ···················· | ···················· | ···················· |
| B | 4156 | hCoV-19/India/DL-NCDC-3279/2020\|EPI_ISL_436434\|202 | ···················· | ···················· | ····················· | ···················· | ···················· | ···················· |
| B | 4157 | hCoV-19/India/DL-NCDC-3394/2020\|EPI_ISL_436435\|202 | ···················· | ···················· | ····················· | ···················· | ···················· | ···················· |
| B | 4158 | hCoV-19/India/DL-NCDC-3400/2020\|EPI_ISL_436436\|202 | ···················· | ···················· | ····················· | ···················· | ···················· | ···················· |
| B | 4159 | hCoV-19/India/DL-NCDC-3831/2020\|EPI_ISL_436437\|202 | ···················· | ···················· | ····················· | ···················· | ···················· | ···················· |

| | | | | | | | |
|---|---|---|---|---|---|---|---|
| B | 4160 hCoV-19/Saudi Arabia/KAUST-Makkah229/2020\|EPI_ISL_ | ···················· | ···················· | ···················· | ···················· | ···················· | ···················· |
| B | 4161 hCoV-19/Saudi Arabia/KAUST-Makkah230/2020\|EPI_ISL_ | ···················· | ···················· | ···················· | ···················· | ···················· | ···················· |
| B | 4162 hCoV-19/Saudi Arabia/KAUST-Makkah231/2020\|EPI_ISL_ | ···················· | ···················· | ···················· | ···················· | ···················· | ···················· |
| B | 4163 hCoV-19/Saudi Arabia/KAUST-Makkah232/2020\|EPI_ISL_ | ···················· | ···················· | ···················· | ···················· | ···················· | ···················· |
| B | 4164 hCoV-19/Saudi Arabia/KAUST-Makkah233/2020\|EPI_ISL_ | ···················· | ···················· | ···················· | ···················· | ···················· | ··A················· |
| B | 4165 hCoV-19/Saudi Arabia/KAUST-Makkah234/2020\|EPI_ISL_ | ···················· | ···················· | ···················· | ···················· | ···················· | ···················· |
| B | 4166 hCoV-19/Saudi Arabia/KAUST-Makkah236/2020\|EPI_ISL_ | ···················· | ·A·················· | ···················· | ···················· | ···················· | ··A················· |
| B | 4167 hCoV-19/Saudi Arabia/KAUST-Makkah237/2020\|EPI_ISL_ | ···················· | ···················· | ···················· | ···················· | ···················· | ···················· |
| B | 4168 hCoV-19/Saudi Arabia/KAUST-Madinah238/2020\|EPI_ISL | ···················· | ···················· | ···················· | ···················· | ···················· | ···················· |
| B | 4169 hCoV-19/Saudi Arabia/KAUST-Madinah248/2020\|EPI_ISL | ···················· | ···················· | ···················· | ···················· | ···················· | ···················· |
| B | 4170 hCoV-19/Saudi Arabia/KAUST-Madinah249/2020\|EPI_ISL | ···················· | ···················· | ···················· | ···················· | ···················· | ···················· |
| B | 4171 hCoV-19/Saudi Arabia/KAUST-Madinah252/2020\|EPI_ISL | ···················· | ·A·················· | ···················· | ···················· | ···················· | ··A················· |
| B | 4172 hCoV-19/Saudi Arabia/KAUST-Madinah253/2020\|EPI_ISL | ···················· | ···················· | ···················· | ···················· | ···················· | ···················· |
| B | 4173 hCoV-19/Saudi Arabia/KAUST-Madinah254/2020\|EPI_ISL | ···················· | ···················· | ···················· | ···················· | ···················· | ···················· |
| B | 4174 hCoV-19/Saudi Arabia/KAUST-Madinah255/2020\|EPI_ISL | ···················· | ·A·················· | ···················· | ···················· | ···················· | ··A················· |
| B | 4175 hCoV-19/Saudi Arabia/KAUST-Madinah256/2020\|EPI_ISL | ···················· | ···················· | ···················· | ···················· | ···················· | ···················· |
| B | 4176 hCoV-19/Saudi Arabia/KAUST-Madinah257/2020\|EPI_ISL | ···················· | ···················· | ···················· | ···················· | ···················· | ···················· |
| B | 4177 hCoV-19/Saudi Arabia/KAUST-Madinah258/2020\|EPI_ISL | ···················· | ···················· | ···················· | ···················· | ···················· | ···················· |
| B | 4178 hCoV-19/Saudi Arabia/KAUST-Madinah260/2020\|EPI_ISL | ···················· | ···················· | ···················· | ···················· | ···················· | ···················· |
| B | 4179 hCoV-19/Saudi Arabia/KAUST-Madinah261/2020\|EPI_ISL | ···················· | ···················· | ···················· | ···················· | ···················· | ···················· |
| B | 4180 hCoV-19/Saudi Arabia/KAUST-Madinah262/2020\|EPI_ISL | ···················· | ···················· | ···················· | ···················· | ···················· | ···················· |
| B | 4181 hCoV-19/Saudi Arabia/KAUST-Madinah263/2020\|EPI_ISL | ···················· | ···················· | ···················· | ···················· | ···················· | ···················· |
| B | 4182 hCoV-19/Saudi Arabia/KAUST-Madinah267/2020\|EPI_ISL | ···················· | ···················· | ···················· | ···················· | ···················· | ···················· |
| B | 4183 hCoV-19/Saudi Arabia/KAUST-Madinah269/2020\|EPI_ISL | ···················· | ···················· | ···················· | ···················· | ···················· | ···················· |
| B | 4184 hCoV-19/Saudi Arabia/KAUST-Madinah270/2020\|EPI_ISL | ···················· | ·A·················· | ···················· | ···················· | ···················· | ···················· |
| B | 4185 hCoV-19/Saudi Arabia/KAUST-Madinah272/2020\|EPI_ISL | ···················· | ···················· | ···················· | ···················· | ···················· | ···················· |
| B | 4186 hCoV-19/Saudi Arabia/KAUST-Jeddah61/2020\|EPI_ISL_4 | ···················· | ···················· | ···················· | ···················· | ···················· | ···················· |
| B | 4187 hCoV-19/Saudi Arabia/KAUST-Jeddah66/2020\|EPI_ISL_4 | ···················· | ···················· | ···················· | ···················· | ···················· | ···················· |
| B | 4188 hCoV-19/Saudi Arabia/KAUST-Jeddah70/2020\|EPI_ISL_4 | ···················· | ···················· | ···················· | ···················· | ···················· | ···················· |
| B | 4189 hCoV-19/Saudi Arabia/KAUST-Jeddah73/2020\|EPI_ISL_4 | ···················· | ···················· | ···················· | ···················· | ···················· | ···················· |
| B | 4190 hCoV-19/Saudi Arabia/KAUST-Jeddah75/2020\|EPI_ISL_4 | ···················· | ···················· | ···················· | ···················· | ···················· | ···················· |
| B | 4191 hCoV-19/Saudi Arabia/KAUST-Jeddah76/2020\|EPI_ISL_4 | ···················· | ···················· | ···················· | ···················· | ···················· | ···················· |
| B | 4192 hCoV-19/Saudi Arabia/KAUST-Makkah90/2020\|EPI_ISL_4 | ···················· | ···················· | ···················· | ···················· | ···················· | ···················· |
| B | 4193 hCoV-19/Saudi Arabia/KAUST-Makkah97/2020\|EPI_ISL_4 | ···················· | ···················· | ···················· | ···················· | ···················· | ···················· |
| B | 4194 hCoV-19/USA/VA-DCLS-0118/2020\|EPI_ISL_437763\|2020- | ···················· | ···················· | ···················· | ···················· | ···················· | ···················· |
| B | 4195 hCoV-19/USA/VA-DCLS-0120/2020\|EPI_ISL_437764\|2020- | ···················· | ···················· | ···················· | ···················· | ···················· | ···················· |
| B | 4196 hCoV-19/USA/VA-DCLS-0126/2020\|EPI_ISL_437765\|2020- | ···················· | ···················· | ···················· | ···················· | ···················· | ···················· |
| B | 4197 hCoV-19/USA/VA-DCLS-0127/2020\|EPI_ISL_437766\|2020- | ···················· | ···················· | ···················· | ···················· | ···················· | ···················· |
| B | 4198 hCoV-19/USA/VA-DCLS-0128/2020\|EPI_ISL_437767\|2020- | ···················· | ···················· | ···················· | ···················· | ···················· | ···················· |
| B | 4199 hCoV-19/USA/VA-DCLS-0129/2020\|EPI_ISL_437768\|2020- | ···················· | ···················· | ···················· | ···················· | ···················· | ···················· |
| B | 4200 hCoV-19/Greece/248_32261/2020\|EPI_ISL_437900\|2020- | ···················· | ···················· | ···················· | ···················· | ···················· | ···················· |
| B | 4201 hCoV-19/Greece/251_32400/2020\|EPI_ISL_437901\|2020- | ···················· | ···················· | ···················· | ···················· | ···················· | ···················· |
| B | 4202 hCoV-19/Greece/153_33892/2020\|EPI_ISL_437902\|2020- | ···················· | ···················· | ···················· | ···················· | ···················· | ···················· |
| B | 4203 hCoV-19/Greece/243_32202/2020\|EPI_ISL_437903\|2020- | ···················· | ···················· | ···················· | ···················· | ···················· | ···················· |
| B | 4204 hCoV-19/Greece/244_32204/2020\|EPI_ISL_437904\|2020- | ···················· | ···················· | ···················· | ···················· | ···················· | ···················· |
| B | 4205 hCoV-19/Greece/246_32206/2020\|EPI_ISL_437905\|2020- | ···················· | ···················· | ···················· | ···················· | ···················· | ···················· |
| B | 4206 hCoV-19/Greece/260_32216/2020\|EPI_ISL_437906\|2020- | ···················· | ···················· | ···················· | ···················· | ···················· | ···················· |
| B | 4207 hCoV-19/Greece/261_32221/2020\|EPI_ISL_437907\|2020- | ···················· | ···················· | ···················· | ···················· | ···················· | ···················· |
| B | 4208 hCoV-19/Greece/264_32497/2020\|EPI_ISL_437908\|2020- | ···················· | ···················· | ···················· | ···················· | ···················· | ···················· |
| B | 4209 hCoV-19/Greece/49_36716/2020\|EPI_ISL_437909\|2020-0 | ···················· | ···················· | ···················· | ···················· | ···················· | ···················· |
| B | 4210 hCoV-19/Greece/52_37266/2020\|EPI_ISL_437910\|2020-0 | ···················· | ···················· | ···················· | ···················· | ···················· | ···················· |
| B | 4211 hCoV-19/Greece/56_37161/2020\|EPI_ISL_437911\|2020-0 | ···················· | ···················· | ···················· | ···················· | ···················· | ···················· |
| B | 4212 hCoV-19/Bangladesh/CHRF-0001/2020\|EPI_ISL_437912\|2 | ···················· | ···················· | ···················· | ···················· | ···················· | ···················· |
| B | 4213 hCoV-19/Austria/CeMM0023/2020\|EPI_ISL_437913\|2020- | ···················· | ···················· | ···················· | ···················· | ···················· | ···················· |
| B | 4214 hCoV-19/Austria/CeMM0024/2020\|EPI_ISL_437914\|2020- | ···················· | ···················· | ···················· | ···················· | ···················· | ···················· |
| B | 4215 hCoV-19/Austria/CeMM0025/2020\|EPI_ISL_437915\|2020- | ···················· | ···················· | ···················· | ···················· | ···················· | ···················· |
| B | 4216 hCoV-19/Austria/CeMM0026/2020\|EPI_ISL_437916\|2020- | ···················· | ···················· | ···················· | ···················· | ···················· | ···················· |
| B | 4217 hCoV-19/Austria/CeMM0028/2020\|EPI_ISL_437917\|2020- | ···················· | ···················· | ···················· | ···················· | ···················· | ···················· |
| B | 4218 hCoV-19/Austria/CeMM0029/2020\|EPI_ISL_437918\|2020- | ···················· | ···················· | ···················· | ···················· | ···················· | ···················· |
| B | 4219 hCoV-19/Austria/CeMM0030/2020\|EPI_ISL_437919\|2020- | ···················· | ···················· | ···················· | ···················· | ···················· | ···················· |
| B | 4220 hCoV-19/Austria/CeMM0031/2020\|EPI_ISL_437920\|2020- | ···················· | ···················· | ···················· | ···················· | ···················· | ···················· |
| B | 4221 hCoV-19/Austria/CeMM0032/2020\|EPI_ISL_437921\|2020- | ···················· | ···················· | ···················· | ···················· | ···················· | ···················· |
| B | 4222 hCoV-19/Austria/CeMM0033/2020\|EPI_ISL_437922\|2020- | ···················· | ···················· | ···················· | ···················· | ···················· | ···················· |
| B | 4223 hCoV-19/England/CAMB-795F4/2020\|EPI_ISL_441102\|202 | ···················· | ···················· | ···················· | ···················· | ···················· | ···················· |
| B | 4224 hCoV-19/England/CAMB-7806D/2020\|EPI_ISL_441103\|202 | ···················· | ···················· | ···················· | ···················· | ···················· | ···················· |
| B | 4225 hCoV-19/England/CAMB-7949D/2020\|EPI_ISL_441104\|202 | ···················· | ···················· | ···················· | ···················· | ···················· | ···················· |
| B | 4226 hCoV-19/England/CAMB-7816A/2020\|EPI_ISL_441107\|202 | ···················· | ···················· | ···················· | ···················· | ···················· | ···················· |
| B | 4227 hCoV-19/England/CAMB-7A39F/2020\|EPI_ISL_441108\|202 | ···················· | ···················· | ···················· | ···················· | ···················· | ···················· |
| B | 4228 hCoV-19/England/CAMB-7814C/2020\|EPI_ISL_441110\|202 | ···················· | ···················· | ···················· | ···················· | ···················· | ···················· |
| B | 4229 hCoV-19/England/CAMB-78B02/2020\|EPI_ISL_441111\|202 | ···················· | ···················· | ···················· | ···················· | ···················· | ···················· |
| B | 4230 hCoV-19/England/CAMB-7815B/2020\|EPI_ISL_441112\|202 | ···················· | ···················· | ···················· | ···················· | ···················· | ···················· |
| B | 4231 hCoV-19/England/CAMB-79460/2020\|EPI_ISL_441114\|202 | ···················· | ···················· | ···················· | ···················· | ···················· | ···················· |
| B | 4232 hCoV-19/England/CAMB-78DB1/2020\|EPI_ISL_441115\|202 | ···················· | ···················· | ···················· | ···················· | ···················· | ···················· |
| B | 4233 hCoV-19/England/CAMB-780A9/2020\|EPI_ISL_441118\|202 | ···················· | ···················· | ···················· | ···················· | ···················· | ···················· |
| B | 4234 hCoV-19/England/CAMB-795B8/2020\|EPI_ISL_441119\|202 | ···················· | ···················· | ···················· | ···················· | ···················· | ···················· |
| B | 4235 hCoV-19/England/CAMB-77F61/2020\|EPI_ISL_441120\|202 | ···················· | ···················· | ···················· | ···················· | ···················· | ···················· |
| B | 4236 hCoV-19/England/CAMB-7B64A/2020\|EPI_ISL_441122\|202 | ···················· | ···················· | ···················· | ···················· | ···················· | ···················· |
| B | 4237 hCoV-19/England/CAMB-78012/2020\|EPI_ISL_441123\|202 | ···················· | ···················· | ···················· | ···················· | ···················· | ···················· |

| | | | | | | | | |
|---|---|---|---|---|---|---|---|---|
| B | 4238 | hCoV-19/England/CAMB-78C87/2020\|EPI_ISL_441124\|202 | ···················· | ···················· | ···················· | ···················· | ···················· | ···················· |
| B | 4239 | hCoV-19/England/CAMB-77E91/2020\|EPI_ISL_441125\|202 | ···················· | ···················· | ···················· | ···················· | ···················· | ···················· |
| B | 4240 | hCoV-19/England/CAMB-791D2/2020\|EPI_ISL_441126\|202 | ···················· | ···················· | ···················· | ···················· | ···················· | ···················· |
| B | 4241 | hCoV-19/England/CAMB-7A2DE/2020\|EPI_ISL_441127\|202 | ···················· | ···················· | ···················· | ···················· | ···················· | ···················· |
| B | 4242 | hCoV-19/England/CAMB-78030/2020\|EPI_ISL_441128\|202 | ···················· | ···················· | ···················· | ···················· | ···················· | ···················· |
| B | 4243 | hCoV-19/England/CAMB-7911E/2020\|EPI_ISL_441130\|202 | ···················· | ···················· | ···················· | ···················· | ···················· | ···················· |
| B | 4244 | hCoV-19/England/CAMB-794F7/2020\|EPI_ISL_441131\|202 | ···················· | ···················· | ···················· | ···················· | ···················· | ···················· |
| B | 4245 | hCoV-19/England/CAMB-7A863/2020\|EPI_ISL_441132\|202 | ···················· | ···················· | ···················· | ···················· | ···················· | ···················· |
| B | 4246 | hCoV-19/England/CAMB-7A739/2020\|EPI_ISL_441135\|202 | ···················· | ···················· | ···················· | ···················· | ···················· | ···················· |
| B | 4247 | hCoV-19/England/CAMB-79178/2020\|EPI_ISL_441136\|202 | ···················· | ···················· | ···················· | ···················· | ···················· | ···················· |
| B | 4248 | hCoV-19/England/CAMB-79855/2020\|EPI_ISL_441137\|202 | ···················· | ···················· | ···················· | ···················· | ···················· | ···················· |
| B | 4249 | hCoV-19/England/CAMB-78C5A/2020\|EPI_ISL_441138\|202 | ···················· | ···················· | ···················· | ···················· | ···················· | ···················· |
| B | 4250 | hCoV-19/England/CAMB-78179/2020\|EPI_ISL_441139\|202 | ···················· | ···················· | ···················· | ···················· | ···················· | ···················· |
| B | 4251 | hCoV-19/England/CAMB-79433/2020\|EPI_ISL_441140\|202 | ···················· | ···················· | ···················· | ···················· | ···················· | ···················· |
| B | 4252 | hCoV-19/England/CAMB-79442/2020\|EPI_ISL_441141\|202 | ···················· | ···················· | ···················· | ···················· | ···················· | ···················· |
| B | 4253 | hCoV-19/England/CAMB-7A6E1/2020\|EPI_ISL_441142\|202 | ···················· | ···················· | ···················· | ···················· | ···················· | ···················· |
| B | 4254 | hCoV-19/England/CAMB-79767/2020\|EPI_ISL_441143\|202 | ···················· | ···················· | ···················· | ···················· | ···················· | ···················· |
| B | 4255 | hCoV-19/England/CAMB-78188/2020\|EPI_ISL_441144\|202 | ···················· | ···················· | ···················· | ···················· | ···················· | ···················· |
| B | 4256 | hCoV-19/England/CAMB-797B2/2020\|EPI_ISL_441145\|202 | ···················· | ···················· | ···················· | ···················· | ···················· | ···················· |
| B | 4257 | hCoV-19/England/CAMB-78003/2020\|EPI_ISL_441146\|202 | ···················· | ···················· | ···················· | ···················· | ···················· | ···················· |
| B | 4258 | hCoV-19/England/CAMB-79415/2020\|EPI_ISL_441147\|202 | ···················· | ···················· | ···················· | ···················· | ···················· | ···················· |
| B | 4259 | hCoV-19/England/CAMB-7970D/2020\|EPI_ISL_441148\|202 | ···················· | ···················· | ···················· | ···················· | ···················· | ···················· |
| B | 4260 | hCoV-19/England/CAMB-78CC3/2020\|EPI_ISL_441150\|202 | ···················· | ···················· | ···················· | ···················· | ···················· | ···················· |
| B | 4261 | hCoV-19/England/CAMB-7809A/2020\|EPI_ISL_441152\|202 | ···················· | ···················· | ···················· | ···················· | ···················· | ···················· |
| B | 4262 | hCoV-19/England/CAMB-7973A/2020\|EPI_ISL_441153\|202 | ···················· | ···················· | ···················· | ···················· | ···················· | ···················· |
| B | 4263 | hCoV-19/England/CAMB-79327/2020\|EPI_ISL_441154\|202 | ···················· | ···················· | ···················· | ···················· | ···················· | ···················· |
| B | 4264 | hCoV-19/England/CAMB-79099/2020\|EPI_ISL_441155\|202 | ···················· | ···················· | ···················· | ···················· | ···················· | ···················· |
| B | 4265 | hCoV-19/England/CAMB-79363/2020\|EPI_ISL_441156\|202 | ···················· | ···················· | ···················· | ···················· | ···················· | ···················· |
| B | 4266 | hCoV-19/England/CAMB-77EEC/2020\|EPI_ISL_441157\|202 | ···················· | ···················· | ···················· | ···················· | ···················· | ···················· |
| B | 4267 | hCoV-19/England/CAMB-78C0F/2020\|EPI_ISL_441158\|202 | ···················· | ···················· | ···················· | ···················· | ···················· | ···················· |
| B | 4268 | hCoV-19/England/CAMB-7811F/2020\|EPI_ISL_441160\|202 | ···················· | ···················· | ···················· | ···················· | ···················· | ···················· |
| B | 4269 | hCoV-19/England/CAMB-78C69/2020\|EPI_ISL_441161\|202 | ···················· | ···················· | ···················· | ···················· | ···················· | ···················· |
| B | 4270 | hCoV-19/England/CAMB-77F8F/2020\|EPI_ISL_441162\|202 | ···················· | ···················· | ···················· | ···················· | ···················· | ···················· |
| B | 4271 | hCoV-19/England/CAMB-7954F/2020\|EPI_ISL_441164\|202 | ···················· | ···················· | ···················· | ···················· | ···················· | ···················· |
| B | 4272 | hCoV-19/England/CAMB-79372/2020\|EPI_ISL_441165\|202 | ···················· | ···················· | ···················· | ···················· | ···················· | ···················· |
| B | 4273 | hCoV-19/England/CAMB-79891/2020\|EPI_ISL_441166\|202 | ···················· | ···················· | ···················· | ···················· | ···················· | ···················· |
| B | 4274 | hCoV-19/England/CAMB-7A49C/2020\|EPI_ISL_441167\|202 | ···················· | ···················· | ···················· | ···················· | ···················· | ···················· |
| B | 4275 | hCoV-19/England/CAMB-7907B/2020\|EPI_ISL_441169\|202 | ···················· | ···················· | ···················· | ···················· | ···················· | ···················· |
| B | 4276 | hCoV-19/England/CAMB-78CB4/2020\|EPI_ISL_441170\|202 | ···················· | ···················· | ···················· | ···················· | ···················· | ···················· |
| B | 4277 | hCoV-19/England/CAMB-79336/2020\|EPI_ISL_441171\|202 | ···················· | ···················· | ···················· | ···················· | ···················· | ···················· |
| B | 4278 | hCoV-19/England/CAMB-7A890/2020\|EPI_ISL_441172\|202 | ···················· | ···················· | ···················· | ···················· | ···················· | ···················· |
| B | 4279 | hCoV-19/England/CAMB-798A0/2020\|EPI_ISL_441173\|202 | ···················· | ···················· | ···················· | ···················· | ···················· | ···················· |
| B | 4280 | hCoV-19/England/CAMB-79284/2020\|EPI_ISL_441174\|202 | ···················· | ···················· | ···················· | ···················· | ···················· | ···················· |
| B | 4281 | hCoV-19/England/CAMB-78F9D/2020\|EPI_ISL_441175\|202 | ···················· | ···················· | ···················· | ···················· | ···················· | ···················· |
| B | 4282 | hCoV-19/England/CAMB-7A757/2020\|EPI_ISL_441176\|202 | ···················· | ·A·················· | ···················· | ···················· | ···················· | ···················· |
| B | 4283 | hCoV-19/England/CAMB-7A70C/2020\|EPI_ISL_441177\|202 | ···················· | ···················· | ···················· | ···················· | ···················· | ···················· |
| B | 4284 | hCoV-19/England/CAMB-7A8EB/2020\|EPI_ISL_441178\|202 | ···················· | ···················· | ···················· | ···················· | ···················· | ···················· |
| B | 4285 | hCoV-19/England/CAMB-797FE/2020\|EPI_ISL_441179\|202 | ···················· | ···················· | ···················· | ···················· | ···················· | ···················· |
| B | 4286 | hCoV-19/England/CAMB-793BE/2020\|EPI_ISL_441180\|202 | ···················· | ···················· | ···················· | ···················· | ···················· | ···················· |
| B | 4287 | hCoV-19/England/CAMB-79521/2020\|EPI_ISL_441183\|202 | ···················· | ···················· | ···················· | ···················· | ···················· | ···················· |
| B | 4288 | hCoV-19/England/CAMB-79749/2020\|EPI_ISL_441187\|202 | ···················· | ···················· | ···················· | ···················· | ···················· | ···················· |
| B | 4289 | hCoV-19/England/CAMB-7B5D4/2020\|EPI_ISL_441188\|202 | ···················· | ···················· | ···················· | ···················· | ···················· | ···················· |
| B | 4290 | hCoV-19/England/SHEF-CC6D1/2020\|EPI_ISL_442413\|202 | ···················· | ···················· | ···················· | ···················· | ···················· | ···················· |
| B | 4291 | hCoV-19/England/SHEF-CC880/2020\|EPI_ISL_442418\|202 | ···················· | ···················· | ···················· | ···················· | ···················· | ···················· |
| B | 4292 | hCoV-19/England/SHEF-CD9C8/2020\|EPI_ISL_442438\|202 | ···················· | ···················· | ···················· | ···················· | ···················· | ···················· |
| B | 4293 | hCoV-19/England/SHEF-CEC19/2020\|EPI_ISL_442448\|202 | ···················· | ···················· | ···················· | ···················· | ···················· | ···················· |
| B | 4294 | hCoV-19/England/SHEF-D01F7/2020\|EPI_ISL_442468\|202 | ···················· | ···················· | ···················· | ···················· | ···················· | ···················· |
| B | 4295 | hCoV-19/England/SHEF-D077D/2020\|EPI_ISL_442472\|202 | ···················· | ···················· | ···················· | ···················· | ···················· | ···················· |
| B | 4296 | hCoV-19/England/SHEF-D0CBA/2020\|EPI_ISL_442478\|202 | ···················· | ···················· | ···················· | ···················· | ···················· | ···················· |
| B | 4297 | hCoV-19/England/SHEF-D0CF6/2020\|EPI_ISL_442479\|202 | ···················· | ···················· | ···················· | ···················· | ···················· | ···················· |
| B | 4298 | hCoV-19/England/SHEF-D0EB4/2020\|EPI_ISL_442481\|202 | ···················· | ···················· | ···················· | ···················· | ···················· | ···················· |
| B | 4299 | hCoV-19/England/SHEF-D12C6/2020\|EPI_ISL_442487\|202 | ···················· | ···················· | ···················· | ···················· | ···················· | ···················· |
| B | 4300 | hCoV-19/England/SHEF-D14DF/2020\|EPI_ISL_442489\|202 | ···················· | ···················· | ···················· | ···················· | ···················· | ···················· |
| B | 4301 | hCoV-19/England/BRIS-12530B/2020\|EPI_ISL_443701\|20 | ···················· | ···················· | ···················· | ···················· | ···················· | ············G······· |
| B | 4302 | hCoV-19/England/BRIS-125365/2020\|EPI_ISL_443702\|20 | ···················· | ···················· | ···················· | ···················· | ···················· | ···················· |
| B | 4303 | hCoV-19/England/BRIS-125040/2020\|EPI_ISL_443704\|20 | ···················· | ···················· | ···················· | ···················· | ···················· | ···················· |
| B | 4304 | hCoV-19/England/BRIS-125B5E/2020\|EPI_ISL_443706\|20 | ···················· | ···················· | ···················· | ···················· | ···················· | ···················· |
| B | 4305 | hCoV-19/England/BRIS-1252D1/2020\|EPI_ISL_443707\|20 | ···················· | ···················· | ···················· | ···················· | ···················· | ···················· |
| B | 4306 | hCoV-19/England/BRIS-124B8C/2020\|EPI_ISL_443709\|20 | ···················· | ···················· | ···················· | ···················· | ···················· | ···················· |
| B | 4307 | hCoV-19/England/BRIS-125A42/2020\|EPI_ISL_443710\|20 | ···················· | ···················· | ···················· | ···················· | ···················· | ···················· |
| B | 4308 | hCoV-19/England/BRIS-125918/2020\|EPI_ISL_443711\|20 | ···················· | ···················· | ···················· | ···················· | ···················· | ···················· |
| B | 4309 | hCoV-19/England/BRIS-124858/2020\|EPI_ISL_443714\|20 | ···················· | ···················· | ···················· | ···················· | ···················· | ···················· |
| B | 4310 | hCoV-19/England/BRIS-125462/2020\|EPI_ISL_443715\|20 | ···················· | ···················· | ···················· | ···················· | ···················· | ···················· |
| B | 4311 | hCoV-19/England/BRIS-125E37/2020\|EPI_ISL_443716\|20 | ···················· | ···················· | ···················· | ···················· | ···················· | ···················· |
| B | 4312 | hCoV-19/England/BRIS-125417/2020\|EPI_ISL_443717\|20 | ···················· | ···················· | ···················· | ···················· | ···················· | ···················· |
| B | 4313 | hCoV-19/England/BRIS-125A60/2020\|EPI_ISL_443718\|20 | ···················· | ···················· | ···················· | ···················· | ···················· | ···················· |
| B | 4314 | hCoV-19/England/BRIS-1248A3/2020\|EPI_ISL_443719\|20 | ···················· | ···················· | ···················· | ···················· | ···················· | ···················· |
| B | 4315 | hCoV-19/England/BRIS-124788/2020\|EPI_ISL_443742\|20 | ···················· | ···················· | ···················· | ···················· | ···················· | ···················· |

| | | | | | | | | | |
|---|---|---|---|---|---|---|---|---|---|
| B | 4316 | hCoV-19/England/BRIS-124D59/2020\|EPI_ISL_443743\|20 | ···················· | ···················· | ···················· | ···················· | ···················· | ···················· | |
| B | 4317 | hCoV-19/England/BRIS-12490A/2020\|EPI_ISL_443744\|20 | ···················· | ···················· | ···················· | ···················· | ···················· | ···················· | |
| B | 4318 | hCoV-19/England/BRIS-124955/2020\|EPI_ISL_443745\|20 | ···················· | ···················· | ···················· | ···················· | ···················· | ···················· | |
| B | 4319 | hCoV-19/England/BRIS-1259DC/2020\|EPI_ISL_443746\|20 | ···················· | ···················· | ···················· | ···················· | ···················· | ···················· | |
| B | 4320 | hCoV-19/England/BRIS-124F17/2020\|EPI_ISL_443747\|20 | ···················· | ···················· | ···················· | ···················· | ···················· | ···················· | |
| B | 4321 | hCoV-19/England/BRIS-125480/2020\|EPI_ISL_443748\|20 | ···················· | ···················· | ···················· | ···················· | ···················· | ···················· | |
| B | 4322 | hCoV-19/England/BRIS-125435/2020\|EPI_ISL_443749\|20 | ···················· | ··········A········· | ···················· | ···················· | ···················· | ···················· | |
| B | 4323 | hCoV-19/England/BRIS-1255F6/2020\|EPI_ISL_443751\|20 | ···················· | ···················· | ···················· | ···················· | ···················· | ···················· | |
| B | 4324 | hCoV-19/England/BRIS-125A9D/2020\|EPI_ISL_443752\|20 | ···················· | ···················· | ···················· | ···················· | ···················· | ···················· | |
| B | 4325 | hCoV-19/England/BRIS-124DE0/2020\|EPI_ISL_443764\|20 | ···················· | ···················· | ···················· | ···················· | ···················· | ···················· | |
| B | 4326 | hCoV-19/England/BRIS-125C3D/2020\|EPI_ISL_443765\|20 | ···················· | ···················· | ···················· | ···················· | ···················· | ···················· | |
| B | 4327 | hCoV-19/England/BRIS-124F08/2020\|EPI_ISL_443768\|20 | ···················· | ···················· | ···················· | ···················· | ···················· | ···················· | |
| B | 4328 | hCoV-19/England/BRIS-1247A6/2020\|EPI_ISL_443769\|20 | ···················· | ···················· | ···················· | ···················· | ···················· | ···················· | |
| B | 4329 | hCoV-19/England/BRIS-125329/2020\|EPI_ISL_443770\|20 | ···················· | ···················· | ···················· | ···················· | ···················· | ···················· | |
| B | 4330 | hCoV-19/England/BRIS-124C6B/2020\|EPI_ISL_443772\|20 | ···················· | ···················· | ···················· | ···················· | ···················· | ···················· | |
| B | 4331 | hCoV-19/England/BRIS-124C7A/2020\|EPI_ISL_443773\|20 | ···················· | ···················· | ···················· | ···················· | ···················· | ···················· | |
| B | 4332 | hCoV-19/England/BRIS-125909/2020\|EPI_ISL_443774\|20 | ···················· | ···················· | ···················· | ···················· | ···················· | ···················· | |
| B | 4333 | hCoV-19/England/BRIS-1255C9/2020\|EPI_ISL_443775\|20 | ···················· | ···················· | ···················· | ···················· | ···················· | ···················· | |
| B | 4334 | hCoV-19/England/BRIS-124A34/2020\|EPI_ISL_443776\|20 | ···················· | ···················· | ···················· | ···················· | ···················· | ···················· | |
| B | 4335 | hCoV-19/England/BRIS-125884/2020\|EPI_ISL_443777\|20 | ···················· | ···················· | ···················· | ···················· | ···················· | ···················· | |
| B | 4336 | hCoV-19/England/BRIS-125D85/2020\|EPI_ISL_443778\|20 | ···················· | ···················· | ···················· | ···················· | ···················· | ···················· | |
| B | 4337 | hCoV-19/England/BRIS-125347/2020\|EPI_ISL_443780\|20 | ···················· | ···················· | ···················· | ···················· | ···················· | ···················· | |
| B | 4338 | hCoV-19/England/BRIS-125BF4/2020\|EPI_ISL_443781\|20 | ···················· | ···················· | ···················· | ···················· | ···················· | ···················· | |
| B | 4339 | hCoV-19/England/BRIS-125541/2020\|EPI_ISL_443782\|20 | ···················· | ···················· | ···················· | ···················· | ···················· | ···················· | |
| B | 4340 | hCoV-19/England/BRIS-1256E4/2020\|EPI_ISL_443783\|20 | ···················· | ···················· | ···················· | ···················· | ···················· | ···················· | |
| B | 4341 | hCoV-19/England/BRIS-125C1F/2020\|EPI_ISL_443784\|20 | ···················· | ···················· | ···················· | ···················· | ···················· | ···················· | |
| B | 4342 | hCoV-19/England/BRIS-125E64/2020\|EPI_ISL_443786\|20 | ···················· | ···················· | ···················· | ···················· | ···················· | ···················· | |
| B | 4343 | hCoV-19/England/BRIS-124AE9/2020\|EPI_ISL_443787\|20 | ···················· | ···················· | ···················· | ···················· | ···················· | ···················· | |
| B | 4344 | hCoV-19/England/BRIS-124CD4/2020\|EPI_ISL_443788\|20 | ···················· | ···················· | ···················· | ···················· | ···················· | ···················· | |
| B | 4345 | hCoV-19/England/BRIS-125B30/2020\|EPI_ISL_443789\|20 | ···················· | ···················· | ···················· | ···················· | ···················· | ···················· | |
| B | 4346 | hCoV-19/England/BRIS-125286/2020\|EPI_ISL_443790\|20 | ···················· | ···················· | ···················· | ···················· | ···················· | ···················· | |
| B | 4347 | hCoV-19/England/BRIS-125CD3/2020\|EPI_ISL_443791\|20 | ···················· | ···················· | ···················· | ···················· | ···················· | ···················· | |
| B | 4348 | hCoV-19/England/BRIS-125C97/2020\|EPI_ISL_443793\|20 | ···················· | ···················· | ···················· | ···················· | ···················· | ···················· | |
| B | 4349 | hCoV-19/England/BRIS-125EEC/2020\|EPI_ISL_443795\|20 | ···················· | ···················· | ···················· | ···················· | ···················· | ···················· | |
| B | 4350 | hCoV-19/England/BRIS-125B12/2020\|EPI_ISL_443796\|20 | ···················· | ···················· | ···················· | ···················· | ···················· | ···················· | |
| B | 4351 | hCoV-19/England/BRIS-124C10/2020\|EPI_ISL_443797\|20 | ···················· | ···················· | ···················· | ···················· | ···················· | ···················· | |
| B | 4352 | hCoV-19/England/BRIS-12574B/2020\|EPI_ISL_443798\|20 | ···················· | ···················· | ···················· | ···················· | ···················· | ···················· | |
| B | 4353 | hCoV-19/USA/NY-NYUMC383/2020\|EPI_ISL_430410\|2020-0 | ···················· | ···················· | ···················· | ···················· | ···················· | ···················· | |
| B | 4354 | hCoV-19/USA/NY-NYUMC384/2020\|EPI_ISL_430411\|2020-0 | ···················· | ···················· | ···················· | ···················· | ···················· | ···················· | |
| B | 4355 | hCoV-19/USA/NY-NYUMC385/2020\|EPI_ISL_430412\|2020-0 | ···················· | ···················· | ···················· | ···················· | ···················· | ···················· | |
| B | 4356 | hCoV-19/USA/NY-NYUMC386/2020\|EPI_ISL_430413\|2020-0 | ···················· | ···················· | ···················· | ···················· | ···················· | ···················· | |
| B | 4357 | hCoV-19/USA/NY-NYUMC387/2020\|EPI_ISL_430414\|2020-0 | ···················· | ···················· | ···················· | ···················· | ···················· | ···················· | |
| B | 4358 | hCoV-19/USA/NY-NYUMC388/2020\|EPI_ISL_430415\|2020-0 | ···················· | ···················· | ···················· | ···················· | ···················· | ···················· | |
| B | 4359 | hCoV-19/USA/NY-NYUMC389/2020\|EPI_ISL_430416\|2020-0 | ···················· | ···················· | ···················· | ···················· | ···················· | ···················· | |
| B | 4360 | hCoV-19/USA/NY-NYUMC390/2020\|EPI_ISL_430417\|2020-0 | ···················· | ···················· | ···················· | ···················· | ···················· | ···················· | |
| B | 4361 | hCoV-19/USA/NY-NYUMC391/2020\|EPI_ISL_430418\|2020-0 | ···················· | ···················· | ···················· | ···················· | ···················· | ···················· | |
| B | 4362 | hCoV-19/USA/NY-NYUMC392/2020\|EPI_ISL_430419\|2020-0 | ···················· | ···················· | ···················· | ···················· | ···················· | ···················· | |
| B | 4363 | hCoV-19/USA/NY-NYUMC393/2020\|EPI_ISL_430420\|2020-0 | ···················· | ···················· | ···················· | ···················· | ···················· | ···················· | |
| B | 4364 | hCoV-19/USA/NY-NYUMC394/2020\|EPI_ISL_430421\|2020-0 | ···················· | ···················· | ···················· | ···················· | ···················· | ···················· | |
| B | 4365 | hCoV-19/USA/NY-NYUMC395/2020\|EPI_ISL_430422\|2020-0 | ···················· | ···················· | ···················· | ···················· | ···················· | ···················· | |
| B | 4366 | hCoV-19/USA/NY-NYUMC396/2020\|EPI_ISL_430423\|2020-0 | ···················· | ···················· | ···················· | ···················· | ···················· | ···················· | |
| B | 4367 | hCoV-19/USA/NY-NYUMC397/2020\|EPI_ISL_430424\|2020-0 | ···················· | ···················· | ···················· | ···················· | ···················· | ···················· | |
| B | 4368 | hCoV-19/USA/NY-NYUMC398/2020\|EPI_ISL_430425\|2020-0 | ···················· | ···················· | ···················· | ···················· | ···················· | ···················· | |
| B | 4369 | hCoV-19/USA/NY-NYUMC399/2020\|EPI_ISL_430426\|2020-0 | ···················· | ···················· | ···················· | ···················· | ···················· | ···················· | |
| B | 4370 | hCoV-19/USA/NY-NYUMC400/2020\|EPI_ISL_430427\|2020-0 | ···················· | ···················· | ···················· | ···················· | ···················· | ···················· | |
| B | 4371 | hCoV-19/USA/NJ-NYUMC401/2020\|EPI_ISL_430428\|2020-0 | ···················· | ···················· | ···················· | ···················· | ···················· | ···················· | |
| B | 4372 | hCoV-19/USA/NY-NYUMC402/2020\|EPI_ISL_430429\|2020-0 | ···················· | ···················· | ···················· | ···················· | ···················· | ···················· | |
| B | 4373 | hCoV-19/USA/NY-NYUMC403/2020\|EPI_ISL_430430\|2020-0 | ···················· | ···················· | ···················· | ···················· | ···················· | ···················· | |
| B | 4374 | hCoV-19/USA/NY-NYUMC404/2020\|EPI_ISL_430431\|2020-0 | ···················· | ···················· | ···················· | ···················· | ···················· | ···················· | |
| B | 4375 | hCoV-19/Malaysia/IMR_WC627/2020\|EPI_ISL_430444\|202 | ···················· | ···················· | ···················· | ···················· | ···················· | ···················· | |
| B | 4376 | hCoV-19/mink/Netherlands/1/2020\|EPI_ISL_431778\|202 | ···················· | ···················· | ···················· | ···················· | ···················· | ···················· | |
| B | 4377 | hCoV-19/Fujian/IM3520014T/2020\|EPI_ISL_431782\|2020 | ···················· | ···················· | ···················· | ···················· | ···················· | ···················· | |
| B | 4378 | hCoV-19/Fujian/IM3520016T/2020\|EPI_ISL_431783\|2020 | ···················· | ···················· | ···················· | ···················· | ···················· | ···················· | |
| B | 4379 | hCoV-19/Fujian/IM3520017T/2020\|EPI_ISL_431784\|2020 | ···················· | ···················· | ···················· | ···················· | ···················· | ···················· | |
| B | 4380 | hCoV-19/Indonesia/JK-EIJK-0141/2020\|EPI_ISL_435281 | ···················· | ···················· | ···················· | ···················· | ···················· | ···················· | |
| B | 4381 | hCoV-19/Indonesia/JK-EIJK-0317/2020\|EPI_ISL_435282 | ···················· | ···················· | ···················· | ···················· | ···················· | ···················· | |
| B | 4382 | hCoV-19/Indonesia/JK-EIJK-2444/2020\|EPI_ISL_435283 | ···················· | ···················· | ···················· | ···················· | ···················· | ···················· | |
| B | 4383 | hCoV-19/Israel/CVL-n6267/2020\|EPI_ISL_435284\|2020- | ···················· | ···················· | ···················· | ···················· | ···················· | ···················· | |
| B | 4384 | hCoV-19/Israel/CVL-n6196/2020\|EPI_ISL_435286\|2020- | ···················· | ···················· | ···················· | ···················· | ···················· | ···················· | |
| B | 4385 | hCoV-19/Israel/CVL-nL19/2020\|EPI_ISL_435289\|2020-0 | ···················· | ···················· | ···················· | ···················· | ···················· | ···················· | |
| B | 4386 | hCoV-19/Israel/CVL-nL6/2020\|EPI_ISL_435291\|2020-03 | ···················· | ···················· | ···················· | ···················· | ···················· | ···················· | |
| B | 4387 | hCoV-19/Israel/CVL-nL33/2020\|EPI_ISL_435292\|2020-0 | ···················· | ···················· | ···················· | ···················· | ···················· | ···················· | |
| B | 4388 | hCoV-19/USA/PA-MGSC52-05/2020\|EPI_ISL_436500\|2020- | ···················· | ···················· | ···················· | ···················· | ···················· | ···················· | |
| B | 4389 | hCoV-19/USA/PA-MGSC53-05/2020\|EPI_ISL_436501\|2020- | ···················· | ···················A | ···················· | ···················· | ···················· | ···················· | 1 |
| B | 4390 | hCoV-19/USA/PA-MGSC55-05/2020\|EPI_ISL_436502\|2020- | ···················· | ···················· | ···················· | ···················· | ···················· | ···················· | |
| B | 4391 | hCoV-19/USA/PA-MGSC56-05/2020\|EPI_ISL_436503\|2020- | ···················· | ···················· | ···················· | ···················· | ···················· | ···················· | |
| B | 4392 | hCoV-19/USA/PA-MGSC58-05/2020\|EPI_ISL_436504\|2020 | ···················· | ···················· | ···················· | ···················· | ···················· | ···················· | |
| B | 4393 | hCoV-19/USA/FL-BPHL-08/2020\|EPI_ISL_436512\|2020-04 | ···················· | ···················· | ···················· | ···················· | ···················· | ···················· | |

| | | | | | | | | |
|---|---|---|---|---|---|---|---|---|
| B | 4394 hCoV-19/USA/FL-BPHL-045/2020\|EPI_ISL_436549\|2020-0 | ···················· | ···················· | ···················· | ···················· | ···················· | ···················· | |
| B | 4395 hCoV-19/USA/WI-UW-270/2020\|EPI_ISL_436564\|2020-03- | ···················· | ···················· | ···················· | ···················· | ···················· | ···················· | |
| B | 4396 hCoV-19/USA/WI-UW-271/2020\|EPI_ISL_436565\|2020-04- | ···················· | ···················· | ···················· | ···················· | ···················· | ···················· | |
| B | 4397 hCoV-19/USA/WI-UW-272/2020\|EPI_ISL_436566\|2020-03- | ···················· | ···················· | ···················· | ···················· | ···················· | ···················· | |
| B | 4398 hCoV-19/USA/WI-UW-273/2020\|EPI_ISL_436567\|2020-03- | ···················· | ···················· | ···················· | ···················· | ···················· | ···················· | |
| B | 4399 hCoV-19/USA/WI-UW-274/2020\|EPI_ISL_436568\|2020-03- | ···················· | ···················· | ···················· | ···················· | ···················· | ···················· | |
| B | 4400 hCoV-19/USA/WI-UW-275/2020\|EPI_ISL_436569\|2020-04- | ···················· | ···················· | ···················· | ···················· | ···················· | ···················· | |
| B | 4401 hCoV-19/USA/WI-UW-276/2020\|EPI_ISL_436570\|2020-04- | ···················· | ···················· | ···················· | ···················· | ···················· | ···················· | |
| B | 4402 hCoV-19/USA/WI-UW-277/2020\|EPI_ISL_436571\|2020-04- | ···················· | ···················· | ···················· | ···················· | ···················· | ···················· | |
| B | 4403 hCoV-19/USA/WI-UW-278/2020\|EPI_ISL_436572\|2020-04- | ···················· | ···················· | ···················· | ···················· | ···················· | ···················· | |
| B | 4404 hCoV-19/USA/WI-UW-279/2020\|EPI_ISL_436573\|2020-04- | ···················· | ···················· | ···················· | ···················· | ···················· | ···················· | |
| B | 4405 hCoV-19/USA/WI-UW-280/2020\|EPI_ISL_436574\|2020-04- | ···················· | ···················· | ···················· | ···················· | ···················· | ···················· | |
| B | 4406 hCoV-19/USA/WI-UW-281/2020\|EPI_ISL_436575\|2020-04- | ···················· | ···················· | ···················· | ···················· | ···················· | ···················· | |
| B | 4407 hCoV-19/USA/WI-UW-282/2020\|EPI_ISL_436576\|2020-04- | ···················· | ···················· | ···················· | ···················· | ···················· | ···················· | |
| B | 4408 hCoV-19/USA/WI-UW-283/2020\|EPI_ISL_436577\|2020-04- | ···················· | ···················· | ···················· | ···················· | ···················· | ···················· | |
| B | 4409 hCoV-19/USA/WI-UW-284/2020\|EPI_ISL_436578\|2020-04- | ···················· | ···················· | ···················· | ···················· | ···················· | ···················· | |
| B | 4410 hCoV-19/USA/WI-UW-285/2020\|EPI_ISL_436579\|2020-04- | ···················· | ···················· | ···················· | ···················· | ···················· | ···················· | |
| B | 4411 hCoV-19/USA/WI-UW-286/2020\|EPI_ISL_436580\|2020-04- | ···················· | ···················· | ···················· | ···················· | ···················· | ···················· | |
| B | 4412 hCoV-19/USA/WI-UW-287/2020\|EPI_ISL_436581\|2020-04- | ···················· | ···················· | ···················· | ···················· | ···················· | ···················· | |
| B | 4413 hCoV-19/USA/WI-UW-288/2020\|EPI_ISL_436582\|2020-04- | ···················· | ···················· | ···················· | ···················· | ···················· | ···················· | |
| B | 4414 hCoV-19/USA/WI-UW-289/2020\|EPI_ISL_436583\|2020-04- | ···················· | ···················· | ···················· | ···················· | ···················· | ···················· | |
| B | 4415 hCoV-19/USA/WI-UW-290/2020\|EPI_ISL_436584\|2020-04- | ···················· | ·····A·············· | ···················· | ···················· | ···················· | ···················· | |
| B | 4416 hCoV-19/USA/WI-UW-291/2020\|EPI_ISL_436585\|2020-04- | ···················· | ···················· | ···················· | ···················· | ···················· | ···················· | |
| B | 4417 hCoV-19/USA/WI-UW-292/2020\|EPI_ISL_436586\|2020-04- | ···················· | ···················· | ···················· | ···················· | ···················· | ···················· | |
| B | 4418 hCoV-19/USA/WI-UW-293/2020\|EPI_ISL_436587\|2020-04- | ···················· | ···················· | ···················· | ···················· | ···················· | ···················· | |
| B | 4419 hCoV-19/USA/WI-UW-294/2020\|EPI_ISL_436588\|2020-04- | ···················· | ···················· | ···················· | ···················· | ···················· | ···················· | |
| B | 4420 hCoV-19/USA/WI-UW-295/2020\|EPI_ISL_436589\|2020-04- | ···················· | ···················· | ···················· | ···················· | ···················· | ···················· | |
| B | 4421 hCoV-19/USA/WI-UW-296/2020\|EPI_ISL_436590\|2020-04- | ···················· | ···················· | ···················· | ···················· | ···················· | ···················· | |
| B | 4422 hCoV-19/USA/WI-UW-297/2020\|EPI_ISL_436591\|2020-04- | ···················· | ·····A·············· | ···················· | ···················· | ···················· | ···················· | |
| B | 4423 hCoV-19/USA/WI-UW-298/2020\|EPI_ISL_436592\|2020-04- | ···················· | ···················· | ···················· | ···················· | ···················· | ···················· | |
| B | 4424 hCoV-19/USA/WI-UW-299/2020\|EPI_ISL_436593\|2020-04- | ···················· | ···················· | ···················· | ···················· | ···················· | ···················· | |
| B | 4425 hCoV-19/USA/WI-UW-300/2020\|EPI_ISL_436594\|2020-04- | ···················· | ·····A·············· | ···················· | ···················· | ···················· | ···················· | |
| B | 4426 hCoV-19/USA/WI-UW-301/2020\|EPI_ISL_436595\|2020-04- | ···················· | ···················A | ···················· | ···················· | ···················· | ···················· | 1 |
| B | 4427 hCoV-19/USA/WI-UW-302/2020\|EPI_ISL_436596\|2020-04- | ···················· | ···················· | ···················· | ···················· | ···················· | ···················· | |
| B | 4428 hCoV-19/USA/WI-UW-303/2020\|EPI_ISL_436597\|2020-04- | ···················· | ·····A·············· | ···················· | ···················· | ···················· | ···················· | |
| B | 4429 hCoV-19/USA/WI-UW-304/2020\|EPI_ISL_436598\|2020-04- | ···················· | ···················· | ···················· | ···················· | ···················· | ···················· | |
| B | 4430 hCoV-19/USA/WI-UW-305/2020\|EPI_ISL_436599\|2020-04- | ···················· | ···················· | ···················· | ···················· | ···················· | ···················· | |
| B | 4431 hCoV-19/USA/VA-DCLS-0169/2020\|EPI_ISL_437802\|2020- | ···················· | ···················· | ···················· | ···················· | ···················· | ···················· | |
| B | 4432 hCoV-19/USA/UN-UW-3982/2020\|EPI_ISL_437803\|2020-03 | ···················· | ···················· | ···················· | ···················· | ···················· | ···················· | |
| B | 4433 hCoV-19/USA/WA-UW-4009/2020\|EPI_ISL_437804\|2020-03 | ···················· | ···················· | ···················· | ···················· | ···················· | ···················· | |
| B | 4434 hCoV-19/USA/WA-UW-4014/2020\|EPI_ISL_437805\|2020-03 | ···················· | ···················· | ···················· | ···················· | ···················· | ···················· | |
| B | 4435 hCoV-19/USA/WA-UW-4015/2020\|EPI_ISL_437806\|2020-03 | ···················· | ···················· | ···················· | ···················· | ···················· | ···················· | |
| B | 4436 hCoV-19/USA/WA-UW-4021/2020\|EPI_ISL_437807\|2020-03 | ···················· | ···················· | ···················· | ···················· | ···················· | ···················· | |
| B | 4437 hCoV-19/USA/WA-UW-4024/2020\|EPI_ISL_437808\|2020-03 | ···················· | ···················· | ···················· | ···················· | ···················· | ···················· | |
| B | 4438 hCoV-19/USA/WA-UW-4026/2020\|EPI_ISL_437809\|2020-03 | ···················· | ···················· | ···················· | ···················· | ···················· | ···················· | |
| B | 4439 hCoV-19/USA/WA-UW-4032/2020\|EPI_ISL_437810\|2020-03 | ···················· | ···················· | ···················· | ···················· | ···················· | ···················· | |
| B | 4440 hCoV-19/USA/WA-UW-4041/2020\|EPI_ISL_437811\|2020-03 | ···················· | ···················· | ···················· | ···················· | ···················· | ···················· | |
| B | 4441 hCoV-19/USA/WA-UW-4046/2020\|EPI_ISL_437812\|2020-03 | ···················· | ···················· | ···················· | ···················· | ···················· | ···················· | |
| B | 4442 hCoV-19/USA/WA-UW-4051/2020\|EPI_ISL_437813\|2020-04 | ···················· | ···················· | ···················· | ···················· | ···················· | ···················· | |
| B | 4443 hCoV-19/USA/WA-UW-4058/2020\|EPI_ISL_437814\|2020-04 | ···················· | ···················· | ···················· | ···················· | ···················· | ···················· | |
| B | 4444 hCoV-19/USA/WA-UW-4061/2020\|EPI_ISL_437815\|2020-04 | ···················· | ···················· | ···················· | ···················· | ···················· | ···················· | |
| B | 4445 hCoV-19/USA/WA-UW-4065/2020\|EPI_ISL_437816\|2020-04 | ···················· | ···················· | ···················· | ···················· | ···················· | ···················· | |
| B | 4446 hCoV-19/USA/WA-UW-4067/2020\|EPI_ISL_437817\|2020-04 | ···················· | ···················· | ···················· | ···················· | ···················· | ···················· | |
| B | 4447 hCoV-19/USA/UN-UW-4089/2020\|EPI_ISL_437818\|2020-04 | ···················· | ···················· | ···················· | ···················· | ···················· | ···················· | |
| B | 4448 hCoV-19/USA/UN-UW-4097/2020\|EPI_ISL_437819\|2020-04 | ···················· | ···················· | ···················· | ···················· | ···················· | ···················· | |
| B | 4449 hCoV-19/USA/WA-UW-4114/2020\|EPI_ISL_437820\|2020-03 | ···················· | ···················· | ···················· | ···················· | ···················· | ···················· | |
| B | 4450 hCoV-19/USA/WA-UW-4210/2020\|EPI_ISL_437821\|2020-03 | ···················· | ···················· | ···················· | ···················· | ···················· | ···················· | |
| B | 4451 hCoV-19/USA/WA-UW-4221/2020\|EPI_ISL_437822\|2020-03 | ···················· | ···················· | ···················· | ···················· | ···················· | ···················· | |
| B | 4452 hCoV-19/USA/WA-UW-4225/2020\|EPI_ISL_437824\|2020-03 | ···················· | ···················· | ···················· | ···················· | ···················· | ···················· | |
| B | 4453 hCoV-19/USA/CT-UW-4237/2020\|EPI_ISL_437825\|2020-03 | ···················· | ···················· | ···················· | ···················· | ···················· | ···················· | |
| B | 4454 hCoV-19/USA/ID-UW-4254/2020\|EPI_ISL_437826\|2020-03 | ···················· | ···················· | ···················· | ···················· | ···················· | ···················· | |
| B | 4455 hCoV-19/USA/WA-UW-4270/2020\|EPI_ISL_437827\|2020-03 | ···················· | ···················· | ···················· | ···················· | ···················· | ···················· | |
| B | 4456 hCoV-19/USA/ID-UW-4308/2020\|EPI_ISL_437828\|2020-03 | ···················· | ···················· | ···················· | ···················· | ···················· | ···················· | |
| B | 4457 hCoV-19/USA/WA-UW-4317/2020\|EPI_ISL_437829\|2020-03 | ···················· | ···················· | ···················· | ···················· | ···················· | ···················· | |
| B | 4458 hCoV-19/USA/ID-UW-6446/2020\|EPI_ISL_437830\|2020-04 | ···················· | ···················· | ···················· | ···················· | ···················· | ···················· | |
| B | 4459 hCoV-19/USA/ID-UW-6448/2020\|EPI_ISL_437831\|2020-04 | ···················· | ···················· | ···················· | ···················· | ···················· | ···················· | |
| B | 4460 hCoV-19/USA/ID-UW-6455/2020\|EPI_ISL_437832\|2020-04 | ···················· | ···················· | ···················· | ···················· | ···················· | ···················· | |
| B | 4461 hCoV-19/USA/ID-UW-6457/2020\|EPI_ISL_437833\|2020-04 | ···················· | ···················· | ···················· | ···················· | ···················· | ···················· | |
| B | 4462 hCoV-19/USA/ID-UW-6466/2020\|EPI_ISL_437835\|2020-04 | ···················· | ···················· | ···················· | ···················· | ···················· | ···················· | |
| B | 4463 hCoV-19/USA/ID-UW-6467/2020\|EPI_ISL_437836\|2020-04 | ···················· | ···················· | ···················· | ···················· | ···················· | ···················· | |
| B | 4464 hCoV-19/USA/ID-UW-6472/2020\|EPI_ISL_437837\|2020-04 | ···················· | ···················· | ···················· | ···················· | ···················· | ···················· | |
| B | 4465 hCoV-19/USA/ID-UW-6473/2020\|EPI_ISL_437838\|2020-04 | ···················· | ···················· | ···················· | ···················· | ···················· | ···················· | |
| B | 4466 hCoV-19/USA/WA-UW-6478/2020\|EPI_ISL_437839\|2020-04 | ···················· | ···················· | ···················· | ···················· | ···················· | ···················· | |
| B | 4467 hCoV-19/USA/WA-UW-6480/2020\|EPI_ISL_437840\|2020-04 | ···················· | ···················· | ···················· | ···················· | ···················· | ···················· | |
| B | 4468 hCoV-19/USA/WA-UW-6481/2020\|EPI_ISL_437841\|2020-04 | ···················· | ···················· | ···················· | ···················· | ···················· | ···················· | |
| B | 4469 hCoV-19/USA/WA-UW-6483/2020\|EPI_ISL_437842\|2020-04 | ···················· | ···················· | ···················· | ···················· | ···················· | ···················· | |
| B | 4470 hCoV-19/USA/WA-UW-6484/2020\|EPI_ISL_437843\|2020-04 | ···················· | ···················· | ···················· | ···················· | ···················· | ···················· | |
| B | 4471 hCoV-19/USA/WA-UW-6485/2020\|EPI_ISL_437844\|2020-04 | ···················· | ···················· | ···················· | ···················· | ···················· | ···················· | |

| | | | | | | | | |
|---|---|---|---|---|---|---|---|---|
| B | 4472 hCoV-19/USA/WA-UW-6486/2020\|EPI_ISL_437845\|2020-04 | .................... | .................... | .................... | .................... | .................... | .................... | |
| B | 4473 hCoV-19/USA/WA-UW-6491/2020\|EPI_ISL_437848\|2020-04 | .................... | .................... | .................... | .................... | .................... | .................... | |
| B | 4474 hCoV-19/USA/WA-UW-6497/2020\|EPI_ISL_437850\|2020-04 | .................... | .................... | .................... | .................... | .................... | .................... | |
| B | 4475 hCoV-19/USA/WA-UW-6501/2020\|EPI_ISL_437851\|2020-04 | .................... | .................... | .................... | .................... | .................... | .................... | |
| B | 4476 hCoV-19/USA/UN-UW-6504/2020\|EPI_ISL_437852\|2020-04 | .................... | .................... | .................... | .................... | .................... | .................... | |
| B | 4477 hCoV-19/USA/WA-UW-6507/2020\|EPI_ISL_437853\|2020-04 | .................... | .................... | .................... | .................... | .................... | .................... | |
| B | 4478 hCoV-19/USA/WA-UW-6514/2020\|EPI_ISL_437854\|2020-04 | .................... | .................... | .................... | .................... | .................... | .................... | |
| B | 4479 hCoV-19/USA/WA-UW-6520/2020\|EPI_ISL_437855\|2020-04 | .................... | .................... | .................... | .................... | .................... | .................... | |
| B | 4480 hCoV-19/USA/WA-UW-6526/2020\|EPI_ISL_437856\|2020-04 | .................... | .................... | .................... | .................... | .................... | .................... | |
| B | 4481 hCoV-19/USA/WA-UW-6527/2020\|EPI_ISL_437857\|2020-04 | .................... | .................... | .................... | .................... | .................... | .................... | |
| B | 4482 hCoV-19/USA/WA-UW-6528/2020\|EPI_ISL_437858\|2020-04 | .................... | .................... | .................... | .................... | .................... | .................... | |
| B | 4483 hCoV-19/USA/WA-UW-6535/2020\|EPI_ISL_437859\|2020-04 | .................... | .................... | .................... | .................... | .................... | .................... | |
| B | 4484 hCoV-19/USA/WA-UW-6546/2020\|EPI_ISL_437860\|2020-04 | .................... | .................... | .................... | .................... | .................... | .................... | |
| B | 4485 hCoV-19/USA/WA-UW-6547/2020\|EPI_ISL_437861\|2020-04 | .................... | .................... | .................... | .................... | .................... | .................... | |
| B | 4486 hCoV-19/USA/WA-UW-6548/2020\|EPI_ISL_437862\|2020-04 | .................... | .................... | .................... | .................... | .................... | .................... | |
| B | 4487 hCoV-19/USA/WA-UW-6559/2020\|EPI_ISL_437863\|2020-04 | .................... | .................... | .................... | .................... | .................... | .................... | |
| B | 4488 hCoV-19/USA/WA-UW-6562/2020\|EPI_ISL_437864\|2020-04 | .................... | .................... | .................... | .................... | .................... | .................... | |
| B | 4489 hCoV-19/USA/CT-UW-6564/2020\|EPI_ISL_437865\|2020-04 | .................... | .................... | .................... | .................... | .................... | .................... | |
| B | 4490 hCoV-19/USA/CT-UW-6565/2020\|EPI_ISL_437866\|2020-04 | .................... | ...A................ | .................... | .................... | .................... | .................... | |
| B | 4491 hCoV-19/USA/CT-UW-6569/2020\|EPI_ISL_437868\|2020-04 | .................... | .................... | .................... | .................... | .................... | .................... | |
| B | 4492 hCoV-19/USA/CT-UW-6571/2020\|EPI_ISL_437869\|2020-04 | .................... | .................... | .................... | .................... | .................... | .................... | |
| B | 4493 hCoV-19/USA/CT-UW-6573/2020\|EPI_ISL_437870\|2020-04 | .................... | ...................A | .................... | .................... | .................... | .................... | 1 |
| B | 4494 hCoV-19/USA/CT-UW-6574/2020\|EPI_ISL_437871\|2020-04 | .................... | .................... | .................... | .................... | .................... | .................... | |
| B | 4495 hCoV-19/USA/CT-UW-6576/2020\|EPI_ISL_437872\|2020-04 | .................... | .................... | .................... | .................... | .................... | .................... | |
| B | 4496 hCoV-19/USA/AK-PHL087/2020\|EPI_ISL_437873\|2020-03- | .................... | .................... | .................... | .................... | .................... | .................... | |
| B | 4497 hCoV-19/Greece/43_35679/2020\|EPI_ISL_437874\|2020-0 | .................... | .................... | .................... | ......T............. | .................... | .................... | |
| B | 4498 hCoV-19/Greece/152_33705/2020\|EPI_ISL_437875\|2020- | .................... | .................... | .................... | .................... | .................... | .................... | |
| B | 4499 hCoV-19/Greece/202_32984/2020\|EPI_ISL_437876\|2020- | .................... | .................... | .................... | .................... | .................... | .................... | |
| B | 4500 hCoV-19/Greece/204_33112/2020\|EPI_ISL_437877\|2020- | .................... | .................... | .................... | .................... | .................... | .................... | |
| B | 4501 hCoV-19/Greece/208_33922/2020\|EPI_ISL_437878\|2020- | .................... | .................... | .................... | .................... | .................... | .................... | |
| B | 4502 hCoV-19/Greece/209_33926/2020\|EPI_ISL_437879\|2020- | .................... | .................... | .................... | .................... | .................... | .................... | |
| B | 4503 hCoV-19/Greece/210_33942/2020\|EPI_ISL_437880\|2020- | .................... | .................... | .................... | .................... | .................... | .................... | |
| B | 4504 hCoV-19/Greece/212_34023/2020\|EPI_ISL_437881\|2020- | .................... | .................... | .................... | .................... | .................... | .................... | |
| B | 4505 hCoV-19/Greece/213_34051/2020\|EPI_ISL_437882\|2020- | .................... | .................... | .................... | .................... | .................... | .................... | |
| B | 4506 hCoV-19/Greece/214_34505/2020\|EPI_ISL_437883\|2020- | .................... | .................... | .................... | .................... | .................... | .................... | |
| B | 4507 hCoV-19/Greece/215_34644/2020\|EPI_ISL_437884\|2020- | .................... | .................... | .................... | .................... | .................... | .................... | |
| B | 4508 hCoV-19/Greece/217_34885/2020\|EPI_ISL_437885\|2020- | .................... | .................... | .................... | .................... | .................... | .................... | |
| B | 4509 hCoV-19/Greece/218_35009/2020\|EPI_ISL_437886\|2020- | .................... | .................... | .................... | .................... | .................... | .................... | |
| B | 4510 hCoV-19/Greece/220_35357/2020\|EPI_ISL_437887\|2020- | .................... | .................... | .................... | .................... | .................... | .................... | |
| B | 4511 hCoV-19/Greece/221_33038/2020\|EPI_ISL_437888\|2020- | .................... | .................... | .................... | .................... | .................... | .................... | |
| B | 4512 hCoV-19/Greece/223_34904/2020\|EPI_ISL_437889\|2020- | .................... | .................... | .................... | .................... | .................... | .................... | |
| B | 4513 hCoV-19/Greece/224_35200/2020\|EPI_ISL_437890\|2020- | .................... | .................... | .................... | .................... | .................... | .................... | |
| B | 4514 hCoV-19/Greece/225_35203/2020\|EPI_ISL_437891\|2020- | .................... | .................... | .................... | .................... | .................... | .................... | |
| B | 4515 hCoV-19/Greece/226_35576/2020\|EPI_ISL_437892\|2020- | .................... | .................... | .................... | .................... | .................... | .................... | |
| B | 4516 hCoV-19/Greece/33_36910/2020\|EPI_ISL_437893\|2020-0 | .................... | .................... | .................... | .................... | .................... | .................... | |
| B | 4517 hCoV-19/Greece/234_31670/2020\|EPI_ISL_437894\|2020- | .................... | .................... | .................... | .................... | .................... | .................... | |
| B | 4518 hCoV-19/Greece/236_31815/2020\|EPI_ISL_437895\|2020- | .................... | .................... | .................... | .................... | .................... | .................... | |
| B | 4519 hCoV-19/Greece/237_31890/2020\|EPI_ISL_437896\|2020- | .................... | .................... | .................... | .................... | .................... | .................... | |
| B | 4520 hCoV-19/Greece/40_36244/2020\|EPI_ISL_437897\|2020-0 | .................... | .................... | .................... | .................... | .................... | .................... | |
| B | 4521 hCoV-19/Greece/41_36861/2020\|EPI_ISL_437898\|2020-0 | .................... | .................... | .................... | .................... | .................... | .................... | |
| B | 4522 hCoV-19/Greece/42_36236/2020\|EPI_ISL_437899\|2020-0 | .................... | .................... | .................... | .................... | .................... | .................... | |
| B | 4523 hCoV-19/Austria/CeMM0250/2020\|EPI_ISL_438080\|2020- | .................... | .................... | .................... | .................... | .................... | .................... | |
| B | 4524 hCoV-19/Austria/CeMM0251/2020\|EPI_ISL_438081\|2020- | .................... | .................... | .................... | .................... | .................... | .................... | |
| B | 4525 hCoV-19/Austria/CeMM0252/2020\|EPI_ISL_438082\|2020- | .................... | .................... | .................... | .................... | .................... | .................... | |
| B | 4526 hCoV-19/Austria/CeMM0254/2020\|EPI_ISL_438083\|2020- | .................... | .................... | .................... | .................... | .................... | .................... | |
| B | 4527 hCoV-19/Austria/CeMM0255/2020\|EPI_ISL_438084\|2020- | .................... | .................... | .................... | .................... | .................... | .................... | |
| B | 4528 hCoV-19/Austria/CeMM0256/2020\|EPI_ISL_438085\|2020- | .................... | .................... | .................... | .................... | .................... | .................... | |
| B | 4529 hCoV-19/Austria/CeMM0257/2020\|EPI_ISL_438086\|2020- | .................... | .................... | .................... | .................... | .................... | .................... | |
| B | 4530 hCoV-19/Austria/CeMM0258/2020\|EPI_ISL_438087\|2020- | .................... | .................... | .................... | .................... | .................... | .................... | |
| B | 4531 hCoV-19/Austria/CeMM0259/2020\|EPI_ISL_438088\|2020- | .................... | .................... | .................... | .................... | .................... | .................... | |
| B | 4532 hCoV-19/Austria/CeMM0260/2020\|EPI_ISL_438089\|2020- | .................... | .................... | .................... | .................... | .................... | .................... | |
| B | 4533 hCoV-19/England/CAMB-7558F/2020\|EPI_ISL_440399\|202 | .................... | .................... | .................... | .................... | .................... | .................... | |
| B | 4534 hCoV-19/England/CAMB-78C96/2020\|EPI_ISL_441070\|202 | .................... | .................... | .................... | .................... | .................... | .................... | |
| B | 4535 hCoV-19/England/CAMB-78C2D/2020\|EPI_ISL_441072\|202 | .................... | .................... | .................... | .................... | .................... | .................... | |
| B | 4536 hCoV-19/England/CAMB-78C4B/2020\|EPI_ISL_441073\|202 | .................... | .................... | .................... | .................... | .................... | .................... | |
| B | 4537 hCoV-19/England/CAMB-7958B/2020\|EPI_ISL_441074\|202 | .................... | .................... | .................... | .................... | .................... | .................... | |
| B | 4538 hCoV-19/England/CAMB-793DC/2020\|EPI_ISL_441075\|202 | .................... | .................... | .................... | .................... | .................... | .................... | |
| B | 4539 hCoV-19/England/CAMB-79916/2020\|EPI_ISL_441076\|202 | .................... | .................... | .................... | .................... | .................... | .................... | |
| B | 4540 hCoV-19/England/CAMB-7A3BD/2020\|EPI_ISL_441077\|202 | .................... | .................... | .................... | .................... | .................... | .................... | |
| B | 4541 hCoV-19/England/CAMB-795C7/2020\|EPI_ISL_441079\|202 | .................... | .................... | .................... | .................... | .................... | .................... | |
| B | 4542 hCoV-19/England/CAMB-7913C/2020\|EPI_ISL_441081\|202 | .................... | .................... | .................... | .................... | .................... | .................... | |
| B | 4543 hCoV-19/England/CAMB-798EC/2020\|EPI_ISL_441082\|202 | .................... | .................... | .................... | .................... | .................... | .................... | |
| B | 4544 hCoV-19/England/CAMB-7B686/2020\|EPI_ISL_441083\|202 | .................... | .................... | .................... | .................... | .................... | .................... | |
| B | 4545 hCoV-19/England/CAMB-78D93/2020\|EPI_ISL_441084\|202 | .................... | .................... | .................... | .................... | .................... | .................... | |
| B | 4546 hCoV-19/England/CAMB-79196/2020\|EPI_ISL_441085\|202 | .................... | .................... | .................... | .................... | .................... | .................... | |
| B | 4547 hCoV-19/England/CAMB-79451/2020\|EPI_ISL_441086\|202 | .................... | .................... | .................... | .................... | .................... | .................... | |
| B | 4548 hCoV-19/England/CAMB-7971C/2020\|EPI_ISL_441088\|202 | .................... | .................... | .................... | .................... | .................... | .................... | |
| B | 4549 hCoV-19/England/CAMB-795A9/2020\|EPI_ISL_441089\|202 | .................... | .................... | .................... | .................... | .................... | .................... | |

| | | | | | | | |
|---|---|---|---|---|---|---|---|
| B | 4550 hCoV-19/England/CAMB-79758/2020\|EPI_ISL_441090\|202 | .................... | .................... | .................... | .................... | .................... | .................... |
| B | 4551 hCoV-19/England/CAMB-793CD/2020\|EPI_ISL_441091\|202 | .................... | .................... | .................... | .................... | .................... | .................... |
| B | 4552 hCoV-19/England/CAMB-7A5C6/2020\|EPI_ISL_441092\|202 | .................... | .................... | .................... | .................... | .................... | .................... |
| B | 4553 hCoV-19/England/CAMB-797EF/2020\|EPI_ISL_441093\|202 | .................... | .................... | .................... | .................... | .................... | .................... |
| B | 4554 hCoV-19/England/CAMB-7A2CF/2020\|EPI_ISL_441095\|202 | .................... | .................... | .................... | .................... | .................... | .................... |
| B | 4555 hCoV-19/England/CAMB-78CF0/2020\|EPI_ISL_441096\|202 | .................... | .................... | .................... | .................... | .................... | .................... |
| B | 4556 hCoV-19/England/CAMB-790D5/2020\|EPI_ISL_441098\|202 | .................... | .................... | .................... | .................... | .................... | .................... |
| B | 4557 hCoV-19/England/CAMB-78F7F/2020\|EPI_ISL_441099\|202 | .................... | .................... | .................... | .................... | .................... | .................... |
| B | 4558 hCoV-19/England/CAMB-78FAC/2020\|EPI_ISL_441100\|202 | .................... | .................... | .................... | .................... | .................... | .................... |
| B | 4559 hCoV-19/England/CAMB-7A5B7/2020\|EPI_ISL_441101\|202 | .................... | .................... | .................... | .................... | .................... | .................... |
| B | 4560 hCoV-19/England/SHEF-CC088/2020\|EPI_ISL_442404\|202 | .................... | .................... | .................... | .................... | .................... | .................... |
| B | 4561 hCoV-19/England/SHEF-CC0D3/2020\|EPI_ISL_442407\|202 | .................... | .................... | .................... | .................... | .................... | .................... |
| B | 4562 hCoV-19/England/SHEF-CD55B/2020\|EPI_ISL_442432\|202 | .................... | .................... | .................... | .................... | .................... | .................... |
| B | 4563 hCoV-19/England/BRIS-124B6E/2020\|EPI_ISL_443720\|20 | .................... | .................... | .................... | .................... | .................... | .................... |
| B | 4564 hCoV-19/England/BRIS-125C88/2020\|EPI_ISL_443726\|20 | .................... | .................... | .................... | .................... | .................... | .................... |
| B | 4565 hCoV-19/England/BRIS-124DFF/2020\|EPI_ISL_443729\|20 | .................... | .................... | .................... | .................... | .................... | .................... |
| B | 4566 hCoV-19/England/BRIS-12575A/2020\|EPI_ISL_443730\|20 | .................... | .................... | .................... | .................... | .................... | .................... |
| B | 4567 hCoV-19/England/BRIS-125110/2020\|EPI_ISL_443733\|20 | .................... | .................... | .................... | .................... | .................... | .................... |
| B | 4568 hCoV-19/England/BRIS-1251A7/2020\|EPI_ISL_443738\|20 | .................... | .................... | .................... | .................... | .................... | .................... |
| B | 4569 hCoV-19/England/BRIS-125338/2020\|EPI_ISL_443739\|20 | .................... | .................... | .................... | .................... | .................... | .................... |
| B | 4570 hCoV-19/England/BRIS-125D3A/2020\|EPI_ISL_443740\|20 | .................... | .................... | .................... | .................... | .................... | .................... |
| B | 4571 hCoV-19/England/BRIS-1247E2/2020\|EPI_ISL_443741\|20 | .................... | .................... | .................... | .................... | .................... | .................... |
| B | 4572 hCoV-19/England/BRIS-1249EC/2020\|EPI_ISL_443757\|20 | .................... | .................... | .................... | .................... | .................... | .................... |
| B | 4573 hCoV-19/England/BRIS-12522C/2020\|EPI_ISL_443758\|20 | .................... | .................... | .................... | .................... | .................... | .................... |
| B | 4574 hCoV-19/England/BRIS-12567B/2020\|EPI_ISL_443759\|20 | .................... | .................... | .................... | .................... | .................... | .................... |
| B | 4575 hCoV-19/England/BRIS-125839/2020\|EPI_ISL_443760\|20 | .................... | .................... | .................... | .................... | .................... | .................... |
| B | 4576 hCoV-19/England/BRIS-125AF7/2020\|EPI_ISL_443763\|20 | .................... | .................... | .................... | .................... | .................... | .................... |
| B | 4577 hCoV-19/USA/MI-MDHHS-SC20230/2020\|EPI_ISL_447200\|2 | .................... | .................... | .................... | .................... | .................... | .................... |
| B | 4578 hCoV-19/USA/MI-MDHHS-SC20231/2020\|EPI_ISL_447201\|2 | .................... | .................... | .................... | .................... | .................... | .................... |
| B | 4579 hCoV-19/USA/MI-MDHHS-SC20234/2020\|EPI_ISL_447204\|2 | .................... | .................... | .................... | .................... | .................... | .................... |
| B | 4580 hCoV-19/USA/MI-MDHHS-SC20235/2020\|EPI_ISL_447205\|2 | .................... | .................... | .................... | .................... | .................... | .................... |
| B | 4581 hCoV-19/USA/MI-MDHHS-SC20236/2020\|EPI_ISL_447206\|2 | .................... | .................... | .................... | .................... | .................... | .................... |
| B | 4582 hCoV-19/USA/MI-MDHHS-SC20237/2020\|EPI_ISL_447207\|2 | .................... | .................... | .................... | .................... | .................... | .................... |
| B | 4583 hCoV-19/USA/MI-MDHHS-SC20239/2020\|EPI_ISL_447209\|2 | .................... | .................... | .................... | .................... | .................... | .................... |
| B | 4584 hCoV-19/USA/MI-MDHHS-SC20240/2020\|EPI_ISL_447210\|2 | .................... | .................... | .................... | .................... | .................... | .................... |
| B | 4585 hCoV-19/USA/MI-MDHHS-SC20241/2020\|EPI_ISL_447211\|2 | .................... | .................... | .................... | .................... | .................... | .................... |
| B | 4586 hCoV-19/USA/MI-MDHHS-SC20242/2020\|EPI_ISL_447212\|2 | .................... | .................... | .................... | .................... | .................... | .................... |
| B | 4587 hCoV-19/USA/MI-MDHHS-SC20245/2020\|EPI_ISL_447215\|2 | .................... | .................... | .................... | .................... | .................... | .................... |
| B | 4588 hCoV-19/USA/MI-MDHHS-SC20246/2020\|EPI_ISL_447216\|2 | .................... | .................... | .................... | .................... | .................... | .................... |
| B | 4589 hCoV-19/USA/MI-MDHHS-SC20248/2020\|EPI_ISL_447218\|2 | .................... | .................... | .................... | .................... | .................... | .................... |
| B | 4590 hCoV-19/USA/MI-MDHHS-SC20249/2020\|EPI_ISL_447219\|2 | .................... | .................... | .................... | .................... | .................... | .................... |
| B | 4591 hCoV-19/USA/MI-MDHHS-SC20250/2020\|EPI_ISL_447220\|2 | .................... | .................... | .................... | .................... | .................... | .................... |
| B | 4592 hCoV-19/USA/MI-MDHHS-SC20251/2020\|EPI_ISL_447221\|2 | .................... | .................... | .................... | .................... | .................... | .................... |
| B | 4593 hCoV-19/USA/MI-MDHHS-SC20254/2020\|EPI_ISL_447224\|2 | .................... | .................... | .................... | .................... | .................... | .................... |
| B | 4594 hCoV-19/USA/MI-MDHHS-SC20258/2020\|EPI_ISL_447228\|2 | .................... | .................... | .................... | .................... | .................... | .................... |
| B | 4595 hCoV-19/USA/MI-MDHHS-SC20259/2020\|EPI_ISL_447229\|2 | .................... | .................... | .................... | .................... | .................... | .................... |
| B | 4596 hCoV-19/DRC/3653/2020\|EPI_ISL_447231\|2020-04-23 | .................... | .................... | .................... | .................... | .................... | .................... |
| B | 4597 hCoV-19/DRC/3659/2020\|EPI_ISL_447232\|2020-04-23 | .................... | .................... | .................... | .................... | .................... | .................... |
| B | 4598 hCoV-19/DRC/3661/2020\|EPI_ISL_447233\|2020-04-23 | .................... | .................... | .................... | .................... | .................... | .................... |
| B | 4599 hCoV-19/DRC/3662/2020\|EPI_ISL_447234\|2020-04-23 | .................... | .................... | .................... | .................... | .................... | .................... |
| B | 4600 hCoV-19/DRC/3664/2020\|EPI_ISL_447235\|2020-04-23 | .................... | .................... | .................... | .................... | .................... | .................... |
| B | 4601 hCoV-19/DRC/3683/2020\|EPI_ISL_447236\|2020-04-24 | .................... | .................... | .................... | .................... | .................... | .................... |
| B | 4602 hCoV-19/DRC/3688/2020\|EPI_ISL_447237\|2020-04-24 | .................... | .................... | .................... | .................... | .................... | .................... |
| B | 4603 hCoV-19/DRC/3768/2020\|EPI_ISL_447239\|2020-04-24 | .................... | .................... | .................... | .................... | .................... | .................... |
| B | 4604 hCoV-19/DRC/3778/2020\|EPI_ISL_447240\|2020-04-24 | .................... | .................... | .................... | .................... | .................... | .................... |
| B | 4605 hCoV-19/DRC/3827/2020\|EPI_ISL_447245\|2020-04-24 | .................... | .................... | .................... | .................... | .................... | .................... |
| B | 4607 hCoV-19/DRC/3837/2020\|EPI_ISL_447248\|2020-04-25 | .................... | .................... | .................... | .................... | .................... | .................... |
| B | 4608 hCoV-19/DRC/3841/2020\|EPI_ISL_447249\|2020-04-25 | .................... | .................... | .................... | .................... | .................... | .................... |
| B | 4609 hCoV-19/USA/CT-UW-6362/2020\|EPI_ISL_430971\|2020-04 | .................... | .................... | .................... | .................... | .................... | .................... |
| B | 4610 hCoV-19/USA/CT-UW-6380/2020\|EPI_ISL_430972\|2020-04 | .................... | .................... | .................... | .................... | .................... | .................... |
| B | 4611 hCoV-19/USA/CT-UW-6385/2020\|EPI_ISL_430973\|2020-04 | .................... | .................... | .................... | .................... | .................... | .................... |
| B | 4612 hCoV-19/USA/WA-UW-6399/2020\|EPI_ISL_430974\|2020-04 | .................... | .................... | .................... | .................... | .................... | .................... |
| B | 4613 hCoV-19/USA/WA-UW-6400/2020\|EPI_ISL_430975\|2020-04 | .................... | .................... | .................... | .................... | .................... | .................... |
| B | 4614 hCoV-19/USA/WA-UW-6403/2020\|EPI_ISL_430976\|2020-04 | .................... | .................... | .................... | .................... | .................... | .................... |
| B | 4615 hCoV-19/USA/WA-UW-6404/2020\|EPI_ISL_430977\|2020-04 | .................... | .................... | .................... | .................... | .................... | .................... |
| B | 4616 hCoV-19/USA/WA-UW-6410/2020\|EPI_ISL_430979\|2020-04 | .................... | .................... | .................... | .................... | .................... | .................... |
| B | 4617 hCoV-19/USA/WA-UW-6417/2020\|EPI_ISL_430980\|2020-04 | .................... | .................... | .................... | .................... | .................... | .................... |
| B | 4618 hCoV-19/Scotland/EDB573/2020\|EPI_ISL_433200\|2020-0 | .................... | .................... | .................... | .................... | .................... | .................... |
| B | 4622 hCoV-19/Luxembourg/LNS2818424/2020\|EPI_ISL_434500\| | .................... | .................... | .................... | .................... | .................... | .................... |
| B | 4623 hCoV-19/Luxembourg/LNS5807524/2020\|EPI_ISL_434502\| | .................... | .................... | .................... | .................... | .................... | .................... |
| B | 4624 hCoV-19/Luxembourg/LNS0364174/2020\|EPI_ISL_434503\| | .................... | .................... | .................... | .................... | .................... | .................... |
| B | 4625 hCoV-19/Luxembourg/LNS2458393/2020\|EPI_ISL_434504\| | .................... | .................... | .................... | .................... | .................... | .................... |
| B | 4626 hCoV-19/Luxembourg/LNS9470500/2020\|EPI_ISL_434505\| | .................... | .................... | .................... | .................... | .................... | .................... |
| B | 4627 hCoV-19/Luxembourg/LNS6690193/2020\|EPI_ISL_434506\| | .................... | .................... | .................... | .................... | .................... | .................... |
| B | 4628 hCoV-19/Luxembourg/LNS9175349/2020\|EPI_ISL_434507\| | .................... | .................... | .................... | .................... | .................... | .................... |
| B | 4629 hCoV-19/Luxembourg/LNS2435702/2020\|EPI_ISL_434511\| | .................... | .................... | .................... | .................... | .................... | .................... |
| B | 4630 hCoV-19/Luxembourg/LNS9272998/2020\|EPI_ISL_434512\| | .................... | .................... | .................... | .................... | .................... | .................... |
| B | 4631 hCoV-19/Luxembourg/LNS2629386/2020\|EPI_ISL_434514\| | .................... | .................... | .................... | .................... | .................... | .................... |

| | | | | | | | | |
|---|---|---|---|---|---|---|---|---|
| B | 4632 | hCoV-19/Luxembourg/LNS5953507/2020\|EPI_ISL_434515\| | ···················· | ···················· | ···················· | ···················· | ···················· | ···················· |
| B | 4633 | hCoV-19/Jordan/SR-084/2020\|EPI_ISL_434516\|2020-03- | ···················· | ···················· | ···················· | ···················· | ···················· | ···················· |
| B | 4636 | hCoV-19/USA/LA-ALSR-0060-NBG/2020\|EPI_ISL_434519\|2 | ···················· | ···················· | ···················· | ···················· | ···················· | ···················· |
| B | 4637 | hCoV-19/USA/LA-ALSR-0070-NBG/2020\|EPI_ISL_434526\|2 | ···················· | ···················· | ···················· | ···················· | ···················· | ···················· |
| B | 4638 | hCoV-19/USA/LA-ALSR-0071-NBG/2020\|EPI_ISL_434527\|2 | ···················· | ···················· | ···················· | ···················· | ···················· | ···················· |
| B | 4639 | hCoV-19/USA/LA-ALSR-0076-NBG/2020\|EPI_ISL_434530\|2 | ···················· | ···················· | ···················· | ···················· | ···················· | ···················· |
| B | 4640 | hCoV-19/Costa Rica/01/2020\|EPI_ISL_434533\|2020-03- | ···················· | ···················· | ···················· | ···················· | ···················· | ···················· |
| B | 4641 | hCoV-19/Wuhan/IVDC-HB-GX02/2019\|EPI_ISL_434534\|201 | ···················· | ···················· | ···················· | ···················· | ···················· | ···················· |
| B | 4642 | hCoV-19/Austria/CeMM0154/2020\|EPI_ISL_438000\|2020- | ···················· | ···················· | ···················· | ···················· | ···················· | ···················· |
| B | 4643 | hCoV-19/Austria/CeMM0155/2020\|EPI_ISL_438001\|2020- | ···················· | ···················· | ···················· | ···················· | ···················· | ···················· |
| B | 4644 | hCoV-19/Austria/CeMM0156/2020\|EPI_ISL_438002\|2020- | ···················· | ···················· | ···················· | ···················· | ···················· | ···················· |
| B | 4645 | hCoV-19/Austria/CeMM0157/2020\|EPI_ISL_438003\|2020- | ···················· | ···················· | ···················· | ···················· | ···················· | ···················· |
| B | 4646 | hCoV-19/Austria/CeMM0158/2020\|EPI_ISL_438004\|2020- | ···················· | ···················· | ···················· | ···················· | ···················· | ···················· |
| B | 4647 | hCoV-19/Austria/CeMM0159/2020\|EPI_ISL_438005\|2020- | ···················· | ···················· | ···················· | ···················· | ···················· | ···················· |
| B | 4648 | hCoV-19/Austria/CeMM0160/2020\|EPI_ISL_438006\|2020- | ···················· | ···················· | ···················· | ···················· | ···················· | ···················· |
| B | 4649 | hCoV-19/Austria/CeMM0161/2020\|EPI_ISL_438007\|2020- | ···················· | ···················· | ···················· | ···················· | ···················· | ···················· |
| B | 4650 | hCoV-19/Austria/CeMM0162/2020\|EPI_ISL_438008\|2020- | ···················· | ···················· | ···················· | ···················· | ···················· | ···················· |
| B | 4651 | hCoV-19/Austria/CeMM0163/2020\|EPI_ISL_438009\|2020- | ···················· | ···················· | ···················· | ···················· | ···················· | ···················· |
| B | 4652 | hCoV-19/Austria/CeMM0165/2020\|EPI_ISL_438010\|2020- | ···················· | ···················· | ···················· | ···················· | ···················· | ···················· |
| B | 4653 | hCoV-19/Austria/CeMM0166/2020\|EPI_ISL_438011\|2020- | ···················· | ···················· | ···················· | ···················· | ···················· | ···················· |
| B | 4654 | hCoV-19/Austria/CeMM0167/2020\|EPI_ISL_438012\|2020- | ···················· | ···················· | ···················· | ···················· | ···················· | ···················· |
| B | 4655 | hCoV-19/Austria/CeMM0168/2020\|EPI_ISL_438013\|2020- | ···················· | ···················· | ···················· | ···················· | ···················· | ···················· |
| B | 4656 | hCoV-19/Austria/CeMM0169/2020\|EPI_ISL_438014\|2020- | ···················· | ···················· | ···················· | ···················· | ···················· | ···················· |
| B | 4657 | hCoV-19/Austria/CeMM0170/2020\|EPI_ISL_438015\|2020- | ···················· | ···················· | ···················· | ···················· | ···················· | ···················· |
| B | 4658 | hCoV-19/Austria/CeMM0171/2020\|EPI_ISL_438016\|2020- | ···················· | ···················· | ···················· | ···················· | ···················· | ···················· |
| B | 4659 | hCoV-19/Austria/CeMM0172/2020\|EPI_ISL_438017\|2020- | ···················· | ···················· | ···················· | ···················· | ···················· | ···················· |
| B | 4660 | hCoV-19/Austria/CeMM0175/2020\|EPI_ISL_438018\|2020- | ···················· | ···················· | ···················· | ···················· | ···················· | ···················· |
| B | 4661 | hCoV-19/Austria/CeMM0176/2020\|EPI_ISL_438019\|2020- | ···················· | ···················· | ···················· | ···················· | ···················· | ···················· |
| B | 4662 | hCoV-19/Austria/CeMM0209/2020\|EPI_ISL_438046\|2020- | ···················· | ···················· | ···················· | ···················· | ···················· | ···················· |
| B | 4663 | hCoV-19/Austria/CeMM0210/2020\|EPI_ISL_438047\|2020- | ···················· | ···················· | ···················· | ···················· | ···················· | ···················· |
| B | 4664 | hCoV-19/Austria/CeMM0212/2020\|EPI_ISL_438048\|2020- | ···················· | ···················· | ···················· | ···················· | ···················· | ···················· |
| B | 4665 | hCoV-19/Austria/CeMM0213/2020\|EPI_ISL_438049\|2020- | ···················· | ···················· | ···················· | ···················· | ···················· | ···················· |
| B | 4666 | hCoV-19/Austria/CeMM0226/2020\|EPI_ISL_438060\|2020- | ···················· | ···················· | ···················· | ···················· | ···················· | ···················· |
| B | 4667 | hCoV-19/Austria/CeMM0228/2020\|EPI_ISL_438061\|2020- | ···················· | ···················· | ···················· | ···················· | ···················· | ···················· |
| B | 4668 | hCoV-19/Austria/CeMM0229/2020\|EPI_ISL_438062\|2020- | ···················· | ···················· | ···················· | ···················· | ···················· | ···················· |
| B | 4669 | hCoV-19/Austria/CeMM0230/2020\|EPI_ISL_438063\|2020- | ···················· | ···················· | ···················· | ···················· | ···················· | ···················· |
| B | 4670 | hCoV-19/Austria/CeMM0232/2020\|EPI_ISL_438064\|2020- | ···················· | ···················· | ···················· | ···················· | ···················· | ···················· |
| B | 4671 | hCoV-19/Austria/CeMM0233/2020\|EPI_ISL_438065\|2020- | ···················· | ···················· | ···················· | ···················· | ···················· | ···················· |
| B | 4672 | hCoV-19/Austria/CeMM0235/2020\|EPI_ISL_438066\|2020- | ···················· | ···················· | ···················· | ···················· | ···················· | ···················· |
| B | 4673 | hCoV-19/Austria/CeMM0237/2020\|EPI_ISL_438067\|2020- | ···················· | ···················· | ···················· | ···················· | ···················· | ···················· |
| B | 4674 | hCoV-19/Austria/CeMM0238/2020\|EPI_ISL_438068\|2020- | ···················· | ···················· | ···················· | ···················· | ···················· | ···················· |
| B | 4675 | hCoV-19/Austria/CeMM0239/2020\|EPI_ISL_438069\|2020- | ···················· | ···················· | ···················· | ···················· | ···················· | ···················· |
| B | 4676 | hCoV-19/Austria/CeMM0240/2020\|EPI_ISL_438070\|2020- | ···················· | ···················· | ···················· | ···················· | ···················· | ···················· |
| B | 4677 | hCoV-19/Austria/CeMM0241/2020\|EPI_ISL_438071\|2020- | ···················· | ···················· | ···················· | ···················· | ···················· | ···················· |
| B | 4678 | hCoV-19/Austria/CeMM0242/2020\|EPI_ISL_438072\|2020- | ···················· | ···················· | ···················· | ···················· | ···················· | ···················· |
| B | 4679 | hCoV-19/Austria/CeMM0243/2020\|EPI_ISL_438073\|2020- | ···················· | ···················· | ···················· | ···················· | ···················· | ···················· |
| B | 4680 | hCoV-19/Austria/CeMM0244/2020\|EPI_ISL_438074\|2020- | ···················· | ···················· | ···················· | ···················· | ···················· | ···················· |
| B | 4681 | hCoV-19/Austria/CeMM0245/2020\|EPI_ISL_438075\|2020- | ···················· | ···················· | ···················· | ···················· | ···················· | ···················· |
| B | 4682 | hCoV-19/Austria/CeMM0246/2020\|EPI_ISL_438076\|2020- | ···················· | ···················· | ···················· | ···················· | ···················· | ···················· |
| B | 4683 | hCoV-19/Austria/CeMM0247/2020\|EPI_ISL_438077\|2020- | ···················· | ···················· | ···················· | ···················· | ···················· | ···················· |
| B | 4684 | hCoV-19/Austria/CeMM0248/2020\|EPI_ISL_438078\|2020- | ···················· | ···················· | ···················· | ···················· | ···················· | ···················· |
| B | 4685 | hCoV-19/Austria/CeMM0249/2020\|EPI_ISL_438079\|2020- | ···················· | ···················· | ···················· | ···················· | ···················· | ···················· |
| B | 4686 | hCoV-19/Austria/CeMM0261/2020\|EPI_ISL_438090\|2020- | ···················· | ···················· | ···················· | ···················· | ···················· | ···················· |
| B | 4687 | hCoV-19/Austria/CeMM0262/2020\|EPI_ISL_438091\|2020- | ···················· | ···················· | ···················· | ···················· | ···················· | ···················· |
| B | 4688 | hCoV-19/Austria/CeMM0263/2020\|EPI_ISL_438092\|2020- | ···················· | ···················· | ···················· | ···················· | ···················· | ···················· |
| B | 4689 | hCoV-19/Austria/CeMM0264/2020\|EPI_ISL_438093\|2020- | ···················· | ···················· | ···················· | ···················· | ···················· | ···················· |
| B | 4690 | hCoV-19/Austria/CeMM0265/2020\|EPI_ISL_438094\|2020- | ···················· | ···················· | ···················· | ···················· | ···················· | ···················· |
| B | 4691 | hCoV-19/Austria/CeMM0266/2020\|EPI_ISL_438095\|2020- | ···················· | ···················· | ···················· | ···················· | ···················· | ···················· |
| B | 4692 | hCoV-19/Austria/CeMM0267/2020\|EPI_ISL_438096\|2020- | ···················· | ···················· | ···················· | ···················· | ···················· | ···················· |
| B | 4693 | hCoV-19/Austria/CeMM0269/2020\|EPI_ISL_438097\|2020- | ···················· | ···················· | ···················· | ···················· | ···················· | ···················· |
| B | 4694 | hCoV-19/Austria/CeMM0271/2020\|EPI_ISL_438098\|2020- | ···················· | ···················· | ···················· | ···················· | ···················· | ···················· |
| B | 4695 | hCoV-19/Austria/CeMM0272/2020\|EPI_ISL_438099\|2020- | ···················· | ···················· | ···················· | ···················· | ···················· | ···················· |
| B | 4696 | hCoV-19/Scotland/EDB3658/2020\|EPI_ISL_439309\|2020- | ···················· | ···················· | ···················· | ···················· | ···················· | ···················· |
| B | 4697 | hCoV-19/Scotland/EDB3673/2020\|EPI_ISL_439310\|2020- | ···················· | ·A·················· | ···················· | ···················· | ···················· | ···················· |
| B | 4698 | hCoV-19/Scotland/EDB3695/2020\|EPI_ISL_439311\|2020- | ···················· | ···················· | ···················· | ···················· | ···················· | ···················· |
| B | 4699 | hCoV-19/Scotland/EDB3701/2020\|EPI_ISL_439312\|2020- | ···················· | ·A·················· | ···················· | ···················· | ···················· | ···················· |
| B | 4700 | hCoV-19/Scotland/EDB3725/2020\|EPI_ISL_439313\|2020- | ···················· | ···················· | ···················· | ···················· | ···················· | ···················· |
| B | 4701 | hCoV-19/Scotland/EDB3739/2020\|EPI_ISL_439314\|2020- | ···················· | ···················· | ···················· | ···················· | ···················· | ···················· |
| B | 4702 | hCoV-19/Scotland/EDB3740/2020\|EPI_ISL_439315\|2020- | ···················· | ···················· | ···················· | ···················· | ···················· | ···················· |
| B | 4703 | hCoV-19/Scotland/EDB3744/2020\|EPI_ISL_439316\|2020- | ···················· | ·A·················· | ···················· | ···················· | ···················· | ···················· |
| B | 4704 | hCoV-19/Scotland/EDB3759/2020\|EPI_ISL_439317\|2020- | ···················· | ·A·················· | ···················· | ···················· | ···················· | ···················· |
| B | 4706 | hCoV-19/Scotland/EDB3764/2020\|EPI_ISL_439319\|2020- | ···················· | ·A·················· | ···················· | ···················· | ···················· | ···················· |
| B | 4707 | hCoV-19/Scotland/EDB3775/2020\|EPI_ISL_439320\|2020- | ···················· | ···················· | ···················· | ···················· | ···················· | ···················· |
| B | 4708 | hCoV-19/Scotland/EDB3781/2020\|EPI_ISL_439322\|2020- | ···················· | ·A·················· | ···················· | ···················· | ···················· | ···················· |
| B | 4709 | hCoV-19/Scotland/EDB3783/2020\|EPI_ISL_439323\|2020- | ···················· | ···················· | ···················· | ···················· | ···················· | ···················· |
| B | 4710 | hCoV-19/Scotland/EDB3790/2020\|EPI_ISL_439324\|2020- | ···················· | ·A·················· | ···················· | ···················· | ···················· | ···················· |
| B | 4711 | hCoV-19/Scotland/EDB3795/2020\|EPI_ISL_439326\|2020- | ···················· | ·A·················· | ···················· | ···················· | ···················· | ···················· |
| B | 4712 | hCoV-19/Scotland/EDB3796/2020\|EPI_ISL_439327\|2020- | ···················· | ·A·················· | ···················· | ···················· | ···················· | ···················· |

| | | | | | | | | |
|---|---|---|---|---|---|---|---|---|
| B | 4713 | hCoV-19/Scotland/EDB3798/2020\|EPI_ISL_439328\|2020- | ···················· | ···················· | ···················· | ···················· | ···················· | ···················· |
| B | 4714 | hCoV-19/Scotland/EDB3801/2020\|EPI_ISL_439329\|2020- | ···················· | ···················· | ···················· | ···················· | ···················· | ···················· |
| B | 4715 | hCoV-19/Scotland/EDB3802/2020\|EPI_ISL_439330\|2020- | ···················· | ···················· | ···················· | ···················· | ···················· | ···················· |
| B | 4716 | hCoV-19/Scotland/EDB3803/2020\|EPI_ISL_439331\|2020- | ···················· | ·A·················· | ···················· | ···················· | ···················· | ···················· |
| B | 4717 | hCoV-19/Scotland/EDB3817/2020\|EPI_ISL_439332\|2020- | ···················· | ···················· | ···················· | ···················· | ···················· | ···················· |
| B | 4718 | hCoV-19/Scotland/EDB3826/2020\|EPI_ISL_439333\|2020- | ···················· | ·A·················· | ···················· | ···················· | ···················· | ···················· |
| B | 4719 | hCoV-19/Scotland/EDB3837/2020\|EPI_ISL_439335\|2020- | ···················· | ···················· | ···················· | ···················· | ···················· | ···················· |
| B | 4720 | hCoV-19/Scotland/EDB3842/2020\|EPI_ISL_439336\|2020- | ···················· | ···················· | ···················· | ···················· | ···················· | ···················· |
| B | 4721 | hCoV-19/Scotland/EDB3846/2020\|EPI_ISL_439337\|2020- | ···················· | ···················· | ···················· | ···················· | ···················· | ···················· |
| B | 4722 | hCoV-19/Scotland/EDB3849/2020\|EPI_ISL_439338\|2020- | ···················· | ·A·················· | ···················· | ···················· | ···················· | ···················· |
| B | 4723 | hCoV-19/Scotland/EDB3856/2020\|EPI_ISL_439340\|2020- | ···················· | ···················· | ···················· | ···················· | ···················· | ···················· |
| B | 4724 | hCoV-19/Scotland/EDB3857/2020\|EPI_ISL_439341\|2020- | ···················· | ···················· | ···················· | ···················· | ···················· | ···················· |
| B | 4725 | hCoV-19/Scotland/EDB3859/2020\|EPI_ISL_439342\|2020- | ···················· | ···················· | ···················· | ···················· | ···················· | ···················· |
| B | 4726 | hCoV-19/Scotland/EDB3862/2020\|EPI_ISL_439343\|2020- | ···················· | ···················· | ···················· | ···················· | ···················· | ···················· |
| B | 4727 | hCoV-19/Scotland/EDB3864/2020\|EPI_ISL_439344\|2020- | ···················· | ·A·················· | ···················· | ···················· | ···················· | ···················· |
| B | 4728 | hCoV-19/Scotland/EDB3865/2020\|EPI_ISL_439345\|2020- | ···················· | ···················· | ···················· | ···················· | ···················· | ···················· |
| B | 4729 | hCoV-19/Scotland/EDB3868/2020\|EPI_ISL_439346\|2020- | ···················· | ···················· | ···················· | ···················· | ···················· | ···················· |
| B | 4730 | hCoV-19/Scotland/EDB3869/2020\|EPI_ISL_439347\|2020- | ···················· | ·A·················· | ···················· | ···················· | ···················· | ···················· |
| B | 4731 | hCoV-19/Scotland/EDB3870/2020\|EPI_ISL_439348\|2020- | ···················· | ···················· | ···················· | ···················· | ···················· | ···················· |
| B | 4732 | hCoV-19/Scotland/EDB3871/2020\|EPI_ISL_439349\|2020- | ···················· | ···················· | ···················· | ···················· | ···················· | ···················· |
| B | 4733 | hCoV-19/England/CAMB-76DC2/2020\|EPI_ISL_439376\|202 | ···················· | ···················· | ···················· | ···················· | ···················· | ···················· |
| B | 4734 | hCoV-19/England/CAMB-77268/2020\|EPI_ISL_439377\|202 | ···················· | ···················· | ···················· | ···················· | ···················· | ···················· |
| B | 4735 | hCoV-19/England/CAMB-77374/2020\|EPI_ISL_439378\|202 | ···················· | ···················· | ···················· | ···················· | ···················· | ···················· |
| B | 4736 | hCoV-19/England/CAMB-76436/2020\|EPI_ISL_439379\|202 | ···················· | ···················· | ···················· | ···················· | ···················· | ···················· |
| B | 4737 | hCoV-19/England/CAMB-773B0/2020\|EPI_ISL_439391\|202 | ···················· | ···················· | ···················· | ···················· | ···················· | ···················· |
| B | 4738 | hCoV-19/England/CAMB-779CD/2020\|EPI_ISL_439393\|202 | ···················· | ···················· | ···················· | ···················· | ···················· | ···················· |
| B | 4739 | hCoV-19/England/CAMB-7773C/2020\|EPI_ISL_439396\|202 | ···················· | ···················· | ···················· | ···················· | ···················· | ···················· |
| B | 4740 | hCoV-19/England/CAMB-770F5/2020\|EPI_ISL_439397\|202 | ···················· | ···················· | ···················· | ···················· | ···················· | ···················· |
| B | 4741 | hCoV-19/England/CAMB-77699/2020\|EPI_ISL_439399\|202 | ···················· | ···················· | ···················· | ···················· | ···················· | ···················· |
| B | 4742 | hCoV-19/England/CAMB-732D5/2020\|EPI_ISL_440300\|202 | ···················· | ···················· | ···················· | ···················· | ···················· | ···················· |
| B | 4743 | hCoV-19/England/CAMB-732F3/2020\|EPI_ISL_440301\|202 | ···················· | ···················· | ···················· | ···················· | ···················· | ···················· |
| B | 4744 | hCoV-19/England/CAMB-740E9/2020\|EPI_ISL_440302\|202 | ···················· | ···················· | ···················· | ···················· | ···················· | ···················· |
| B | 4745 | hCoV-19/England/CAMB-74016/2020\|EPI_ISL_440303\|202 | ···················· | ···················· | ···················· | ···················· | ···················· | ···················· |
| B | 4746 | hCoV-19/England/CAMB-73FCF/2020\|EPI_ISL_440304\|202 | ···················· | ···················· | ···················· | ···················· | ···················· | ···················· |
| B | 4747 | hCoV-19/England/CAMB-740F8/2020\|EPI_ISL_440305\|202 | ···················· | ···················· | ···················· | ···················· | ···················· | ···················· |
| B | 4748 | hCoV-19/England/CAMB-740CB/2020\|EPI_ISL_440306\|202 | ···················· | ···················· | ···················· | ···················· | ···················· | ···················· |
| B | 4749 | hCoV-19/England/CAMB-7418C/2020\|EPI_ISL_440307\|202 | ···················· | ···················· | ···················· | ···················· | ···················· | ···················· |
| B | 4750 | hCoV-19/England/CAMB-73F92/2020\|EPI_ISL_440309\|202 | ···················· | ···················· | ···················· | ···················· | ···················· | ···················· |
| B | 4751 | hCoV-19/England/CAMB-7423E/2020\|EPI_ISL_440310\|202 | ···················· | ·············A······ | ···················· | ···················· | ···················· | ···················· |
| B | 4752 | hCoV-19/England/CAMB-73E59/2020\|EPI_ISL_440311\|202 | ···················· | ···················· | ···················· | ···················· | ···················· | ···················· |
| B | 4753 | hCoV-19/England/CAMB-758C2/2020\|EPI_ISL_440313\|202 | ···················· | ···················· | ···················· | ···················· | ···················· | ···················· |
| B | 4754 | hCoV-19/England/CAMB-73F1A/2020\|EPI_ISL_440314\|202 | ···················· | ·············A······ | ···················· | ···················· | ···················· | ···················· |
| B | 4755 | hCoV-19/England/CAMB-73FA1/2020\|EPI_ISL_440315\|202 | ···················· | ···················· | ···················· | ···················· | ···················· | ···················· |
| B | 4756 | hCoV-19/England/CAMB-73F65/2020\|EPI_ISL_440318\|202 | ···················· | ···················· | ···················· | ···················· | ···················· | ···················· |
| B | 4757 | hCoV-19/England/CAMB-74210/2020\|EPI_ISL_440319\|202 | ···················· | ···················· | ···················· | ···················· | ···················· | ···················· |
| B | 4758 | hCoV-19/England/CAMB-73F38/2020\|EPI_ISL_440321\|202 | ···················· | ·············A······ | ···················· | ···················· | ···················· | ···················· |
| B | 4759 | hCoV-19/England/CAMB-73EB3/2020\|EPI_ISL_440323\|202 | ···················· | ···················· | ···················· | ···················· | ···················· | ···················· |
| B | 4760 | hCoV-19/England/CAMB-7419B/2020\|EPI_ISL_440328\|202 | ···················· | ···················· | ···················· | ···················· | ···················· | ···················· |
| B | 4761 | hCoV-19/England/CAMB-740DA/2020\|EPI_ISL_440329\|202 | ···················· | ···················· | ···················· | ···················· | ···················· | ···················· |
| B | 4762 | hCoV-19/England/CAMB-73E1D/2020\|EPI_ISL_440330\|202 | ···················· | ···················· | ···················· | ···················· | ···················· | ···················· |
| B | 4763 | hCoV-19/England/CAMB-75859/2020\|EPI_ISL_440354\|202 | ···················· | ···················· | ···················· | ···················· | ···················· | ···················· |
| B | 4764 | hCoV-19/England/CAMB-758E0/2020\|EPI_ISL_440361\|202 | ···················· | ···················· | ···················· | ···················· | ···················· | ···················· |
| B | 4765 | hCoV-19/England/CAMB-741AA/2020\|EPI_ISL_440364\|202 | ···················· | ···················· | ···················· | ···················· | ···················· | ···················· |
| B | 4766 | hCoV-19/England/CAMB-751F4/2020\|EPI_ISL_440376\|202 | ···················· | ···················· | ···················· | ···················· | ···················· | ···················· |
| B | 4767 | hCoV-19/England/CAMB-753B2/2020\|EPI_ISL_440378\|202 | ···················· | ···················· | ···················· | ···················· | ···················· | ···················· |
| B | 4768 | hCoV-19/England/CAMB-75051/2020\|EPI_ISL_440379\|202 | ···················· | ···················· | ···················· | ···················· | ···················· | ···················· |
| B | 4769 | hCoV-19/England/CAMB-74FBF/2020\|EPI_ISL_440380\|202 | ···················· | ···················· | ···················· | ···················· | ···················· | ···················· |
| B | 4770 | hCoV-19/England/CAMB-753D0/2020\|EPI_ISL_440381\|202 | ···················· | ···················· | ···················· | ···················· | ···················· | ···················· |
| B | 4771 | hCoV-19/England/CAMB-75367/2020\|EPI_ISL_440382\|202 | ···················· | ···················· | ···················· | ···················· | ···················· | ···················· |
| B | 4772 | hCoV-19/England/CAMB-751A9/2020\|EPI_ISL_440384\|202 | ···················· | ···················· | ···················· | ···················· | ···················· | ···················· |
| B | 4773 | hCoV-19/England/CAMB-7508E/2020\|EPI_ISL_440386\|202 | ···················· | ···················· | ···················· | ···················· | ···················· | ···················· |
| B | 4774 | hCoV-19/England/CAMB-7531C/2020\|EPI_ISL_440387\|202 | ···················· | ···················· | ···················· | ···················· | ···················· | ···················· |
| B | 4775 | hCoV-19/England/CAMB-75288/2020\|EPI_ISL_440389\|202 | ···················· | ···················· | ···················· | ···················· | ···················· | ···················· |
| B | 4776 | hCoV-19/England/CAMB-750BB/2020\|EPI_ISL_440391\|202 | ···················· | ···················· | ···················· | ···················· | ···················· | ···················· |
| B | 4777 | hCoV-19/England/CAMB-7509D/2020\|EPI_ISL_440393\|202 | ···················· | ···················· | ···················· | ···················· | ···················· | ···················· |
| B | 4778 | hCoV-19/England/CAMB-752C4/2020\|EPI_ISL_440395\|202 | ···················· | ···················· | ···················· | ···················· | ···················· | ···················· |
| B | 4779 | hCoV-19/England/CAMB-751E5/2020\|EPI_ISL_440396\|202 | ···················· | ···················· | ···················· | ···················· | ···················· | ···················· |
| B | 4780 | hCoV-19/England/CAMB-75279/2020\|EPI_ISL_440397\|202 | ···················· | ···················· | ···················· | ···················· | ···················· | ···················· |
| B | 4781 | hCoV-19/England/CAMB-7C649/2020\|EPI_ISL_441621\|202 | ···················· | ···················· | ···················· | ···················· | ···················· | ···················· |
| B | 4782 | hCoV-19/England/CAMB-7CE6F/2020\|EPI_ISL_441623\|202 | ···················· | ···················· | ···················· | ···················· | ···················· | ···················· |
| B | 4783 | hCoV-19/England/CAMB-7DA88/2020\|EPI_ISL_441624\|202 | ···················· | ···················· | ···················· | ···················· | ···················· | ···················· |
| B | 4784 | hCoV-19/England/CAMB-7DEE6/2020\|EPI_ISL_441626\|202 | ···················· | ···················· | ···················· | ···················· | ···················· | ···················· |
| B | 4785 | hCoV-19/England/CAMB-7DEC8/2020\|EPI_ISL_441627\|202 | ···················· | ···················· | ···················· | ···················· | ···················· | ···················· |
| B | 4786 | hCoV-19/England/CAMB-7C6C1/2020\|EPI_ISL_441628\|202 | ···················· | ···················· | ···················· | ···················· | ···················· | ···················· |
| B | 4787 | hCoV-19/England/CAMB-7DD07/2020\|EPI_ISL_441629\|202 | ···················· | ···················· | ···················· | ···················· | ···················· | ···················· |
| B | 4788 | hCoV-19/England/CAMB-7B99C/2020\|EPI_ISL_441630\|202 | ···················· | ···················· | ···················· | ···················· | ···················· | ···················· |
| B | 4789 | hCoV-19/England/CAMB-7B9D8/2020\|EPI_ISL_441631\|202 | ···················· | ···················· | ···················· | ···················· | ···················· | ···················· |
| B | 4790 | hCoV-19/England/CAMB-7D3F6/2020\|EPI_ISL_441632\|202 | ···················· | ···················· | ···················· | ···················· | ···················· | ···················· |

B 4791 hCoV-19/England/CAMB-7DF89/2020|EPI_ISL_441633|202 ···················· ···················· ···················· ···················· ···················· ····················
B 4792 hCoV-19/England/CAMB-7D87F/2020|EPI_ISL_441634|202 ···················· ···················· ···················· ···················· ···················· ····················
B 4793 hCoV-19/England/CAMB-7B835/2020|EPI_ISL_441635|202 ···················· ···················· ···················· ···················· ···················· ····················
B 4794 hCoV-19/England/CAMB-7C6A3/2020|EPI_ISL_441636|202 ···················· ···················· ···················· ···················· ···················· ····················
B 4795 hCoV-19/England/CAMB-7D9F4/2020|EPI_ISL_441637|202 ···················· ···················· ···················· ···················· ···················· ····················
B 4796 hCoV-19/England/CAMB-7CD35/2020|EPI_ISL_441638|202 ···················· ···················· ···················· ···················· ···················· ····················
B 4797 hCoV-19/England/CAMB-7DC64/2020|EPI_ISL_441639|202 ···················· ···················· ···················· ···················· ···················· ····················
B 4798 hCoV-19/England/CAMB-7CCB0/2020|EPI_ISL_441640|202 ···················· ···················· ···················· ···················· ···················· ····················
B 4799 hCoV-19/England/CAMB-7C53D/2020|EPI_ISL_441641|202 ···················· ···················· ···················· ···················· ···················· ····················
B 4800 hCoV-19/England/CAMB-7CDEA/2020|EPI_ISL_441642|202 ···················· ···················· ···················· ···················· ···················· ····················
B 4801 hCoV-19/England/CAMB-7B9F6/2020|EPI_ISL_441643|202 ···················· ···················· ···················· ···················· ···················· ····················
B 4802 hCoV-19/England/CAMB-7CDAE/2020|EPI_ISL_441644|202 ···················· ···················· ···················· ···················· ···················· ····················
B 4803 hCoV-19/England/CAMB-7CB68/2020|EPI_ISL_441645|202 ···················· ···················· ···················· ···················· ···················· ····················
B 4804 hCoV-19/England/CAMB-7CCCF/2020|EPI_ISL_441647|202 ···················· ···················· ···················· ···················· ···················· ····················
B 4805 hCoV-19/England/CAMB-7BAB7/2020|EPI_ISL_441648|202 ···················· ···················· ···················· ···················· ···················· ····················
B 4806 hCoV-19/England/CAMB-7D323/2020|EPI_ISL_441649|202 ···················· ···················· ···················· ···················· ···················· ····················
B 4807 hCoV-19/England/CAMB-7B747/2020|EPI_ISL_441650|202 ···················· ···················· ···················· ···················· ···················· ····················
B 4808 hCoV-19/England/CAMB-7BA4E/2020|EPI_ISL_441651|202 ···················· ···················· ···················· ···················· ···················· ····················
B 4809 hCoV-19/England/CAMB-7CBE0/2020|EPI_ISL_441653|202 ···················· ···················· ···················· ···················· ···················· ····················
B 4810 hCoV-19/England/CAMB-7B6E0/2020|EPI_ISL_441654|202 ···················· ···················· ···················· ···················· ···················· ····················
B 4811 hCoV-19/England/CAMB-7C4A9/2020|EPI_ISL_441656|202 ···················· ···················· ···················· ···················· ···················· ····················
B 4812 hCoV-19/England/CAMB-7D772/2020|EPI_ISL_441657|202 ···················· ···················· ···················· ···················· ···················· ····················
B 4813 hCoV-19/England/CAMB-7B96F/2020|EPI_ISL_441658|202 ···················· ···················· ···················· ···················· ···················· ····················
B 4814 hCoV-19/Northern Ireland/NIRE-FA85C/2020|EPI_ISL_4 ···················· ···················· ···················· ···················· ···················· ····················
B 4815 hCoV-19/England/CAMB-7D8F7/2020|EPI_ISL_441660|202 ···················· ···················· ···················· ···················· ···················· ····················
B 4816 hCoV-19/Northern Ireland/NIRE-FAA1A/2020|EPI_ISL_4 ···················· ···················· ···················· ···················· ···················· ····················
B 4817 hCoV-19/England/CAMB-7DAE2/2020|EPI_ISL_441687|202 ···················· ···················· ···················· ···················· ···················· ····················
B 4818 hCoV-19/England/CAMB-7D9E5/2020|EPI_ISL_441688|202 ···················· ···················· ···················· ···················· ···················· ····················
B 4819 hCoV-19/Northern Ireland/NIRE-FAA56/2020|EPI_ISL_4 ···················· ···················· ···················· ···················· ···················· ····················
B 4820 hCoV-19/Northern Ireland/NIRE-FAA38/2020|EPI_ISL_4 ···················· ···················· ···················· ···················· ···················· ····················
B 4821 hCoV-19/England/CAMB-7DA2E/2020|EPI_ISL_441692|202 ···················· ···················· ···················· ···················· ···················· ····················
B 4822 hCoV-19/Northern Ireland/NIRE-FA986/2020|EPI_ISL_4 ···················· ···················· ···················· ···················· ···················· ····················
B 4823 hCoV-19/Northern Ireland/NIRE-FAA65/2020|EPI_ISL_4 ···················· ···················· ···················· ···················· ···················· ····················
B 4824 hCoV-19/England/CAMB-7D5B4/2020|EPI_ISL_441695|202 ···················· ···················· ···················· ···················· ···················· ····················
B 4825 hCoV-19/England/CAMB-81526/2020|EPI_ISL_442952|202 ···················· ···················· ···················· ···················· ···················· ····················
B 4826 hCoV-19/England/CAMB-80DE3/2020|EPI_ISL_442953|202 ···················· ···················· ···················· ···················· ···················· ····················
B 4827 hCoV-19/England/CAMB-80CB9/2020|EPI_ISL_442954|202 ···················· ···················· ···················· ···················· ···················· ····················
B 4828 hCoV-19/England/CAMB-7EE21/2020|EPI_ISL_442956|202 ···················· ···················· ···················· ···················· ···················· ····················
B 4829 hCoV-19/England/CAMB-7F4B5/2020|EPI_ISL_442957|202 ···················· ···················· ···················· ···················· ···················· ····················
B 4830 hCoV-19/England/CAMB-809B2/2020|EPI_ISL_442959|202 ···················· ···················· ···················· ···················· ···················· ····················
B 4831 hCoV-19/England/CAMB-81580/2020|EPI_ISL_442961|202 ···················· ···················· ···················· ···················· ···················· ····················
B 4832 hCoV-19/England/CAMB-7EB84/2020|EPI_ISL_442962|202 ···················· ···················· ···················· ···················· ···················· ····················
B 4833 hCoV-19/England/CAMB-80CD7/2020|EPI_ISL_442964|202 ···················· ···················· ···················· ···················· ···················· ····················
B 4834 hCoV-19/England/CAMB-7F312/2020|EPI_ISL_442965|202 ···················· ···················· ···················· ···················· ···················· ····················
B 4835 hCoV-19/England/CAMB-80800/2020|EPI_ISL_442966|202 ···················· ···················· ···················· ···················· ···················· ····················
B 4836 hCoV-19/England/CAMB-7F2E8/2020|EPI_ISL_442967|202 ···················· ···················· ···················· ···················· ···················· ····················
B 4837 hCoV-19/England/CAMB-7F4D3/2020|EPI_ISL_442968|202 ···················· ···················· ···················· ···················· ···················· ····················
B 4838 hCoV-19/England/CAMB-7EB48/2020|EPI_ISL_442969|202 ···················· ···················· ···················· ···················· ···················· ····················
B 4839 hCoV-19/England/CAMB-7F075/2020|EPI_ISL_442970|202 ···················· ···················· ···················· ···················· ···················· ····················
B 4840 hCoV-19/England/CAMB-7F39A/2020|EPI_ISL_442973|202 ···················· ···················· ···················· ···················· ···················· ····················
B 4841 hCoV-19/England/CAMB-7EFD3/2020|EPI_ISL_442974|202 ···················· ···················· ···················· ···················· ···················· ····················
B 4842 hCoV-19/England/CAMB-7EEE5/2020|EPI_ISL_442975|202 ···················· ···················· ···················· ···················· ···················· ····················
B 4843 hCoV-19/England/CAMB-8127A/2020|EPI_ISL_442976|202 ···················· ···················· ···················· ···················· ···················· ····················
B 4844 hCoV-19/England/CAMB-7F34F/2020|EPI_ISL_442977|202 ···················· ···················· ···················· ···················· ···················· ····················
B 4845 hCoV-19/England/CAMB-7F057/2020|EPI_ISL_442978|202 ···················· ···················· ···················· ···················· ···················· ····················
B 4846 hCoV-19/England/CAMB-80E0E/2020|EPI_ISL_442979|202 ···················· ···················· ···················· ···················· ···················· ····················
B 4847 hCoV-19/England/CAMB-81544/2020|EPI_ISL_442980|202 ···················· ···················· ···················· ···················· ···················· ····················
B 4848 hCoV-19/England/CAMB-80A19/2020|EPI_ISL_442981|202 ···················· ···················· ···················· ···················· ···················· ····················
B 4849 hCoV-19/England/CAMB-7F172/2020|EPI_ISL_442982|202 ···················· ···················· ···················· ···················· ···················· ····················
B 4850 hCoV-19/England/CAMB-80C6E/2020|EPI_ISL_442983|202 ···················· ···················· ···················· ···················· ···················· ····················
B 4851 hCoV-19/England/CAMB-7EE6D/2020|EPI_ISL_442984|202 ···················· ···················· ···················· ···················· ···················· ····················
B 4852 hCoV-19/England/CAMB-7F3D6/2020|EPI_ISL_442985|202 ···················· ···················· ···················· ···················· ···················· ····················
B 4853 hCoV-19/England/CAMB-7F0FD/2020|EPI_ISL_442987|202 ···················· ···················· ···················· ···················· ···················· ····················
B 4854 hCoV-19/England/CAMB-81623/2020|EPI_ISL_442988|202 ···················· ···················· ···················· ···················· ···················· ····················
B 4855 hCoV-19/England/CAMB-81201/2020|EPI_ISL_442989|202 ···················· ···················· ···················· ···················· ···················· ····················
B 4856 hCoV-19/England/CAMB-80DF2/2020|EPI_ISL_442991|202 ···················· ···················· ···················· ···················· ···················· ····················
B 4857 hCoV-19/England/CAMB-80BDA/2020|EPI_ISL_442993|202 ···················· ·············A······ ···················· ···················· ···················· ····················
B 4858 hCoV-19/Wales/PHWC-318E0/2020|EPI_ISL_446501|2020- ···················· ···················· ···················· ···················· ···················· ····················
B 4859 hCoV-19/Wales/PHWC-31929/2020|EPI_ISL_446505|2020- ···················· ···················· ···················· ···················· ···················· ····················
B 4860 hCoV-19/Greece/34_36284/2020|EPI_ISL_447832|2020-0 ···················· ···················· ···················· ···················· ···················· ····················
B 4861 hCoV-19/Greece/50_36277/2020|EPI_ISL_447833|2020-0 ···················· ···················· ···················· ···················· ···················· ····················
B 4862 hCoV-19/Greece/55_36015/2020|EPI_ISL_447834|2020-0 ···················· ···················· ···················· ···················· ···················· ····················
B 4863 hCoV-19/Greece/54_37163/2020|EPI_ISL_447835|2020-0 ···················· ···················· ···················· ···················· ···················· ····················
B 4864 hCoV-19/Greece/145_34726/2020|EPI_ISL_447836|2020- ···················· ···················· ···················· ···················· ···················· ····················
B 4865 hCoV-19/USA/WA-UW-5205/2020|EPI_ISL_430900|2020-04 ···················· ···················· ···················· ···················· ···················· ····················
B 4866 hCoV-19/USA/WA-UW-5359/2020|EPI_ISL_430901|2020-04 ···················· ···················· ···················· ···················· ···················· ····················
B 4867 hCoV-19/USA/WA-UW-5364/2020|EPI_ISL_430902|2020-04 ···················· ···················· ···················· ···················· ···················· ····················
B 4868 hCoV-19/USA/WA-UW-5371/2020|EPI_ISL_430904|2020-04 ···················· ···················· ···················· ···················· ···················· ····················

| | | | | | | | | |
|---|---|---|---|---|---|---|---|---|
| B | 4869 | hCoV-19/USA/WA-UW-5397/2020\|EPI_ISL_430905\|2020-04 | .................... | .................... | .................... | .................... | .................... | .................... |
| B | 4870 | hCoV-19/USA/WA-UW-5400/2020\|EPI_ISL_430906\|2020-04 | .................... | .................... | .................... | .................... | .................... | .................... |
| B | 4871 | hCoV-19/USA/CT-UW-5595/2020\|EPI_ISL_430907\|2020-04 | .................... | .................... | .................... | .................... | .................... | .................... |
| B | 4872 | hCoV-19/USA/CT-UW-5599/2020\|EPI_ISL_430908\|2020-04 | .................... | .................... | .................... | .................... | .................... | .................... |
| B | 4873 | hCoV-19/USA/UN-UW-5605/2020\|EPI_ISL_430909\|2020-04 | .................... | .................... | .................... | .................... | .................... | .................... |
| B | 4874 | hCoV-19/USA/UN-UW-5614/2020\|EPI_ISL_430910\|2020-04 | .................... | .................... | .................... | .................... | .................... | .................... |
| B | 4875 | hCoV-19/USA/WA-UW-5617/2020\|EPI_ISL_430911\|2020-04 | .................... | .................... | .................... | .................... | .................... | .................... |
| B | 4876 | hCoV-19/USA/WA-UW-5629/2020\|EPI_ISL_430912\|2020-04 | .................... | .................... | .................... | .................... | .................... | .................... |
| B | 4877 | hCoV-19/USA/WA-UW-5732/2020\|EPI_ISL_430914\|2020-04 | .................... | .................... | .................... | .................... | .................... | .................... |
| B | 4878 | hCoV-19/USA/WA-UW-5737/2020\|EPI_ISL_430915\|2020-04 | .................... | .................... | .................... | .................... | .................... | .................... |
| B | 4879 | hCoV-19/USA/UN-UW-5742/2020\|EPI_ISL_430916\|2020-04 | .................... | .................... | .................... | .................... | .................... | .................... |
| B | 4880 | hCoV-19/USA/CT-UW-5749/2020\|EPI_ISL_430917\|2020-04 | .................... | .................... | .................... | .................... | .................... | .................... |
| B | 4881 | hCoV-19/USA/WA-UW-5750/2020\|EPI_ISL_430918\|2020-04 | .................... | .................... | .................... | .................... | .................... | .................... |
| B | 4882 | hCoV-19/USA/CT-UW-5754/2020\|EPI_ISL_430919\|2020-04 | .................... | .................... | .................... | .................... | .................... | .................... |
| B | 4883 | hCoV-19/USA/WA-UW-5756/2020\|EPI_ISL_430920\|2020-04 | .................... | .................... | .................... | .................... | .................... | .................... |
| B | 4884 | hCoV-19/USA/CT-UW-5757/2020\|EPI_ISL_430921\|2020-04 | .................... | .................... | .................... | .................... | .................... | .................... |
| B | 4885 | hCoV-19/USA/CT-UW-5763/2020\|EPI_ISL_430922\|2020-04 | .................... | .................... | .................... | .................... | .................... | .................... |
| B | 4886 | hCoV-19/USA/CT-UW-5764/2020\|EPI_ISL_430923\|2020-04 | .................... | .................... | .................... | .................... | .................... | .................... |
| B | 4887 | hCoV-19/USA/CT-UW-5770/2020\|EPI_ISL_430925\|2020-04 | .................... | .................... | .................... | .................... | .................... | .................... |
| B | 4888 | hCoV-19/USA/CT-UW-5772/2020\|EPI_ISL_430926\|2020-04 | .................... | .................... | .................... | .................... | .................... | .................... |
| B | 4889 | hCoV-19/USA/CT-UW-5773/2020\|EPI_ISL_430927\|2020-04 | .................... | .................... | .................... | .................... | .................... | .................... |
| B | 4890 | hCoV-19/USA/WA-UW-5879/2020\|EPI_ISL_430928\|2020-04 | .................... | .................... | .................... | .................... | .................... | .................... |
| B | 4891 | hCoV-19/USA/WA-UW-5884/2020\|EPI_ISL_430929\|2020-04 | .................... | .................... | .................... | .................... | .................... | .................... |
| B | 4892 | hCoV-19/USA/OR-UW-4410/2020\|EPI_ISL_430950\|2020-03 | .................... | .................... | .................... | .................... | .................... | .................... |
| B | 4893 | hCoV-19/USA/WA-UW-4413/2020\|EPI_ISL_430951\|2020-03 | .................... | .................... | .................... | .................... | .................... | .................... |
| B | 4894 | hCoV-19/USA/ID-UW-4418/2020\|EPI_ISL_430952\|2020-03 | .................... | .................... | .................... | .................... | .................... | .................... |
| B | 4895 | hCoV-19/USA/WA-UW-6185/2020\|EPI_ISL_430953\|2020-04 | .................... | .................... | .................... | .................... | .................... | .................... |
| B | 4896 | hCoV-19/USA/WA-UW-6186/2020\|EPI_ISL_430954\|2020-04 | .................... | .................... | .................... | .................... | .................... | .................... |
| B | 4897 | hCoV-19/USA/WA-UW-6211/2020\|EPI_ISL_430955\|2020-04 | .................... | .................... | .................... | .................... | .................... | .................... |
| B | 4898 | hCoV-19/USA/WA-UW-6240/2020\|EPI_ISL_430957\|2020-04 | .................... | .................... | .................... | .................... | .................... | .................... |
| B | 4899 | hCoV-19/USA/WA-UW-6241/2020\|EPI_ISL_430958\|2020-04 | .................... | .................... | .................... | .................... | .................... | .................... |
| B | 4900 | hCoV-19/USA/WA-UW-6243/2020\|EPI_ISL_430959\|2020-04 | .................... | .................... | .................... | .................... | .................... | .................... |
| B | 4901 | hCoV-19/USA/CT-UW-6259/2020\|EPI_ISL_430960\|2020-04 | .................... | .................... | .................... | .................... | .................... | .................... |
| B | 4902 | hCoV-19/USA/CT-UW-6270/2020\|EPI_ISL_430961\|2020-04 | .................... | .................... | .................... | .................... | .................... | .................... |
| B | 4903 | hCoV-19/USA/CT-UW-6271/2020\|EPI_ISL_430962\|2020-04 | .................... | .................... | .................... | .................... | .................... | .................... |
| B | 4904 | hCoV-19/USA/WA-UW-6302/2020\|EPI_ISL_430964\|2020-04 | .................... | .................... | .................... | .................... | .................... | .................... |
| B | 4905 | hCoV-19/USA/CT-UW-6314/2020\|EPI_ISL_430965\|2020-04 | .................... | .................... | .................... | .................... | .................... | .................... |
| B | 4906 | hCoV-19/USA/WA-UW-6327/2020\|EPI_ISL_430967\|2020-04 | .................... | .................... | .................... | .................... | .................... | .................... |
| B | 4907 | hCoV-19/USA/WA-UW-6331/2020\|EPI_ISL_430968\|2020-04 | .................... | .................... | .................... | .................... | .................... | .................... |
| B | 4908 | hCoV-19/USA/WA-UW-6335/2020\|EPI_ISL_430969\|2020-04 | .................... | .................... | .................... | .................... | .................... | .................... |
| B | 4909 | hCoV-19/Scotland/EDB2062/2020\|EPI_ISL_433170\|2020- | .................... | .................... | .................... | .................... | .................... | .................... |
| B | 4910 | hCoV-19/Scotland/EDB2064/2020\|EPI_ISL_433171\|2020- | .................... | .................... | .................... | .................... | .................... | .................... |
| B | 4911 | hCoV-19/Scotland/EDB2065/2020\|EPI_ISL_433172\|2020- | .................... | .................... | .................... | .................... | .................... | .................... |
| B | 4912 | hCoV-19/Scotland/EDB2066/2020\|EPI_ISL_433173\|2020- | .................... | .A.................. | .................... | .................... | .................... | .................... |
| B | 4913 | hCoV-19/Scotland/EDB2071/2020\|EPI_ISL_433174\|2020- | .................... | .................... | .................... | .................... | .................... | .................... |
| B | 4914 | hCoV-19/Scotland/EDB2075/2020\|EPI_ISL_433175\|2020- | .................... | .................... | .................... | .................... | .................... | .................... |
| B | 4915 | hCoV-19/Scotland/EDB2077/2020\|EPI_ISL_433176\|2020- | .................... | .................... | .................... | .................... | .................... | .................... |
| B | 4916 | hCoV-19/Scotland/EDB2078/2020\|EPI_ISL_433177\|2020- | .................... | .................... | .................... | .................... | .................... | .................... |
| B | 4917 | hCoV-19/Scotland/EDB2079/2020\|EPI_ISL_433178\|2020- | .................... | .A.................. | .................... | .................... | .................... | .................... |
| B | 4918 | hCoV-19/Scotland/EDB2081/2020\|EPI_ISL_433180\|2020- | .................... | .................... | .................... | .................... | .................... | .................... |
| B | 4919 | hCoV-19/Scotland/EDB2082/2020\|EPI_ISL_433181\|2020- | .................... | .................... | .................... | .................... | .................... | .................... |
| B | 4920 | hCoV-19/Scotland/EDB2083/2020\|EPI_ISL_433182\|2020- | .................... | .A.................. | .................... | .................... | .................... | .................... |
| B | 4921 | hCoV-19/Scotland/EDB2084/2020\|EPI_ISL_433183\|2020- | .................... | .................... | .................... | .................... | .................... | .................... |
| B | 4922 | hCoV-19/Scotland/EDB2085/2020\|EPI_ISL_433184\|2020- | .................... | .A.................. | .................... | .................... | .................... | .................... |
| B | 4923 | hCoV-19/Scotland/EDB2086/2020\|EPI_ISL_433185\|2020- | .................... | .................... | .................... | .................... | .................... | .................... |
| B | 4924 | hCoV-19/Scotland/EDB2087/2020\|EPI_ISL_433186\|2020- | .................... | .................... | .................... | .................... | .................... | .................... |
| B | 4925 | hCoV-19/Scotland/EDB2088/2020\|EPI_ISL_433187\|2020- | .................... | .................... | .................... | .................... | .................... | .................... |
| B | 4926 | hCoV-19/Scotland/EDB2089/2020\|EPI_ISL_433188\|2020- | .................... | .A.................. | .................... | .................... | .................... | .................... |
| B | 4927 | hCoV-19/Scotland/EDB2092/2020\|EPI_ISL_433190\|2020- | .................... | .................... | .................... | .................... | .................... | .................... |
| B | 4928 | hCoV-19/Scotland/EDB2094/2020\|EPI_ISL_433192\|2020- | .................... | .................... | .................... | .................... | .................... | .................... |
| B | 4929 | hCoV-19/Scotland/EDB2095/2020\|EPI_ISL_433193\|2020- | .................... | .................... | .................... | .................... | .................... | .................... |
| B | 4930 | hCoV-19/Scotland/EDB2096/2020\|EPI_ISL_433194\|2020- | .................... | .................... | .................... | .................... | .................... | .................... |
| B | 4931 | hCoV-19/Scotland/EDB2097/2020\|EPI_ISL_433195\|2020- | .................... | .................... | .................... | .................... | .................... | .................... |
| B | 4932 | hCoV-19/Scotland/EDB2098/2020\|EPI_ISL_433196\|2020- | .................... | .A.................. | .................... | .................... | .................... | .................... |
| B | 4933 | hCoV-19/Scotland/EDB2099/2020\|EPI_ISL_433197\|2020- | .................... | .................... | .................... | .................... | .................... | .................... |
| B | 4934 | hCoV-19/Scotland/EDB328/2020\|EPI_ISL_433198\|2020-0 | .................... | .................... | .................... | .................... | .................... | .................... |
| B | 4935 | hCoV-19/Austria/CeMM0177/2020\|EPI_ISL_438020\|2020- | .................... | .................... | .................... | .................... | .................... | .................... |
| B | 4936 | hCoV-19/Austria/CeMM0178/2020\|EPI_ISL_438021\|2020- | .................... | .................... | .................... | .................... | .................... | .................... |
| B | 4937 | hCoV-19/Austria/CeMM0180/2020\|EPI_ISL_438022\|2020- | .................... | .................... | .................... | .................... | .................... | .................... |
| B | 4938 | hCoV-19/Austria/CeMM0181/2020\|EPI_ISL_438023\|2020- | .................... | .................... | .................... | .................... | .................... | .................... |
| B | 4939 | hCoV-19/Austria/CeMM0182/2020\|EPI_ISL_438024\|2020- | .................... | .................... | .................... | .................... | .................... | .................... |
| B | 4940 | hCoV-19/Austria/CeMM0183/2020\|EPI_ISL_438025\|2020- | .................... | .................... | .................... | .................... | .................... | .................... |
| B | 4941 | hCoV-19/Austria/CeMM0184/2020\|EPI_ISL_438026\|2020- | .................... | .................... | .................... | .................... | .................... | .................... |
| B | 4942 | hCoV-19/Austria/CeMM0185/2020\|EPI_ISL_438027\|2020- | .................... | .................... | .................... | .................... | .................... | .................... |
| B | 4943 | hCoV-19/Austria/CeMM0186/2020\|EPI_ISL_438028\|2020- | .................... | .................... | .................... | .................... | .................... | .................... |
| B | 4944 | hCoV-19/Austria/CeMM0188/2020\|EPI_ISL_438029\|2020- | .................... | .................... | .................... | .................... | .................... | .................... |
| B | 4945 | hCoV-19/Austria/CeMM0189/2020\|EPI_ISL_438030\|2020- | .................... | .................... | .................... | .................... | .................... | .................... |
| B | 4946 | hCoV-19/Austria/CeMM0190/2020\|EPI_ISL_438031\|2020- | .................... | .................... | .................... | .................... | .................... | .................... |

| | | | | | | | | |
|---|---|---|---|---|---|---|---|---|
| B | 4947 | hCoV-19/Austria/CeMM0191/2020\|EPI_ISL_438032\|2020- | ···················· | ···················· | ····················· | ···················· | ···················· | ···················· |
| B | 4948 | hCoV-19/Austria/CeMM0192/2020\|EPI_ISL_438033\|2020- | ···················· | ···················· | ····················· | ···················· | ···················· | ···················· |
| B | 4949 | hCoV-19/Austria/CeMM0193/2020\|EPI_ISL_438034\|2020- | ···················· | ···················· | ····················· | ···················· | ···················· | ···················· |
| B | 4950 | hCoV-19/Austria/CeMM0194/2020\|EPI_ISL_438035\|2020- | ···················· | ···················· | ····················· | ···················· | ···················· | ···················· |
| B | 4951 | hCoV-19/Austria/CeMM0195/2020\|EPI_ISL_438036\|2020- | ···················· | ···················· | ····················· | ···················· | ···················· | ···················· |
| B | 4952 | hCoV-19/Austria/CeMM0196/2020\|EPI_ISL_438037\|2020- | ···················· | ···················· | ····················· | ···················· | ···················· | ···················· |
| B | 4953 | hCoV-19/Austria/CeMM0197/2020\|EPI_ISL_438038\|2020- | ···················· | ···················· | ····················· | ···················· | ···················· | ···················· |
| B | 4954 | hCoV-19/Austria/CeMM0198/2020\|EPI_ISL_438039\|2020- | ···················· | ···················· | ····················· | ···················· | ···················· | ···················· |
| B | 4955 | hCoV-19/Austria/CeMM0199/2020\|EPI_ISL_438040\|2020- | ···················· | ···················· | ····················· | ···················· | ···················· | ···················· |
| B | 4956 | hCoV-19/Austria/CeMM0200/2020\|EPI_ISL_438041\|2020- | ···················· | ···················· | ····················· | ···················· | ···················· | ···················· |
| B | 4957 | hCoV-19/Austria/CeMM0201/2020\|EPI_ISL_438042\|2020- | ···················· | ···················· | ····················· | ···················· | ···················· | ···················· |
| B | 4958 | hCoV-19/Austria/CeMM0204/2020\|EPI_ISL_438043\|2020- | ···················· | ···················· | ····················· | ···················· | ···················· | ···················· |
| B | 4959 | hCoV-19/Austria/CeMM0205/2020\|EPI_ISL_438044\|2020- | ···················· | ···················· | ····················· | ···················· | ···················· | ···················· |
| B | 4960 | hCoV-19/Austria/CeMM0206/2020\|EPI_ISL_438045\|2020- | ···················· | ···················· | ····················· | ···················· | ···················· | ···················· |
| B | 4961 | hCoV-19/Austria/CeMM0216/2020\|EPI_ISL_438050\|2020- | ···················· | ···················· | ····················· | ···················· | ···················· | ···················· |
| B | 4962 | hCoV-19/Austria/CeMM0217/2020\|EPI_ISL_438051\|2020- | ···················· | ···················· | ····················· | ···················· | ···················· | ···················· |
| B | 4963 | hCoV-19/Austria/CeMM0218/2020\|EPI_ISL_438052\|2020- | ···················· | ···················· | ····················· | ···················· | ···················· | ···················· |
| B | 4964 | hCoV-19/Austria/CeMM0219/2020\|EPI_ISL_438053\|2020- | ···················· | ···················· | ····················· | ···················· | ···················· | ···················· |
| B | 4965 | hCoV-19/Austria/CeMM0220/2020\|EPI_ISL_438054\|2020- | ···················· | ···················· | ····················· | ···················· | ···················· | ···················· |
| B | 4966 | hCoV-19/Austria/CeMM0221/2020\|EPI_ISL_438055\|2020- | ···················· | ···················· | ····················· | ···················· | ···················· | ···················· |
| B | 4967 | hCoV-19/Austria/CeMM0222/2020\|EPI_ISL_438056\|2020- | ···················· | ···················· | ····················· | ···················· | ···················· | ···················· |
| B | 4968 | hCoV-19/Austria/CeMM0223/2020\|EPI_ISL_438057\|2020- | ···················· | ···················· | ····················· | ···················· | ···················· | ···················· |
| B | 4969 | hCoV-19/Austria/CeMM0224/2020\|EPI_ISL_438058\|2020- | ···················· | ···················· | ····················· | ···················· | ···················· | ···················· |
| B | 4970 | hCoV-19/Austria/CeMM0225/2020\|EPI_ISL_438059\|2020- | ···················· | ···················· | ····················· | ···················· | ···················· | ···················· |
| B | 4971 | hCoV-19/Scotland/EDB3657/2020\|EPI_ISL_439308\|2020- | ···················· | ·A·················· | ····················· | ···················· | ···················· | ···················· |
| B | 4972 | hCoV-19/Scotland/EDB3872/2020\|EPI_ISL_439350\|2020- | ···················· | ···················· | ····················· | ···················· | ···················· | ···················· |
| B | 4973 | hCoV-19/Scotland/EDB3873/2020\|EPI_ISL_439351\|2020- | ···················· | ·A·················· | ····················· | ···················· | ···················· | ···················· |
| B | 4974 | hCoV-19/Scotland/EDB3879/2020\|EPI_ISL_439352\|2020- | ···················· | ···················· | ····················· | ···················· | ···················· | ···················· |
| B | 4975 | hCoV-19/Scotland/EDB3882/2020\|EPI_ISL_439353\|2020- | ···················· | ···················· | ····················· | ···················· | ···················· | ···················· |
| B | 4976 | hCoV-19/Scotland/EDB3887/2020\|EPI_ISL_439354\|2020- | ···················· | ···················· | ····················· | ···················· | ···················· | ···················· |
| B | 4977 | hCoV-19/Scotland/EDB3890/2020\|EPI_ISL_439355\|2020- | ···················· | ···················· | ····················· | ···················· | ···················· | ···················· |
| B | 4978 | hCoV-19/Scotland/EDB3892/2020\|EPI_ISL_439357\|2020- | ···················· | ···················· | ····················· | ···················· | ···················· | ···················· |
| B | 4979 | hCoV-19/Scotland/EDB3894/2020\|EPI_ISL_439358\|2020- | ···················· | ···················· | ····················· | ···················· | ···················· | ···················· |
| B | 4980 | hCoV-19/Scotland/EDB3895/2020\|EPI_ISL_439359\|2020- | ···················· | ···················· | ····················· | ···················· | ···················· | ···················· |
| B | 4981 | hCoV-19/Scotland/EDB3897/2020\|EPI_ISL_439360\|2020- | ···················· | ···················· | ····················· | ···················· | ···················· | ···················· |
| B | 4982 | hCoV-19/Scotland/EDB3899/2020\|EPI_ISL_439361\|2020- | ···················· | ···················· | ····················· | ···················· | ···················· | ···················· |
| B | 4983 | hCoV-19/Scotland/EDB3903/2020\|EPI_ISL_439363\|2020- | ···················· | ···················· | ····················· | ···················· | ···················· | ···················· |
| B | 4984 | hCoV-19/Scotland/EDB3906/2020\|EPI_ISL_439364\|2020- | ···················· | ···················· | ····················· | ···················· | ···················· | ···················· |
| B | 4985 | hCoV-19/Scotland/EDB3914/2020\|EPI_ISL_439365\|2020- | ···················· | ···················· | ····················· | ···················· | ···················· | ···················· |
| B | 4986 | hCoV-19/Scotland/EDB3918/2020\|EPI_ISL_439366\|2020- | ·T·················· | ···················· | ····················· | ···················· | ···················· | ···················· |
| B | 4987 | hCoV-19/Scotland/EDB3919/2020\|EPI_ISL_439367\|2020- | ···················· | ···················· | ····················· | ···················· | ···················· | ···················· |
| B | 4988 | hCoV-19/England/CAMB-7766C/2020\|EPI_ISL_439368\|202 | ···················· | ···················· | ····················· | ···················· | ···················· | ···················· |
| B | 4989 | hCoV-19/England/CAMB-77839/2020\|EPI_ISL_439369\|202 | ···················· | ···················· | ····················· | ···················· | ···················· | ···················· |
| B | 4990 | hCoV-19/England/CAMB-7716B/2020\|EPI_ISL_439371\|202 | ···················· | ···················· | ····················· | ···················· | ···················· | ···················· |
| B | 4991 | hCoV-19/England/CAMB-77A33/2020\|EPI_ISL_439372\|202 | ···················· | ···················· | ····················· | ···················· | ···················· | ···················· |
| B | 4992 | hCoV-19/England/CAMB-77B12/2020\|EPI_ISL_439373\|202 | ···················· | ···················· | ····················· | ···················· | ···················· | ···················· |
| B | 4993 | hCoV-19/England/CAMB-776E4/2020\|EPI_ISL_439374\|202 | ···················· | ···················· | ····················· | ···················· | ···················· | ···················· |
| B | 4994 | hCoV-19/England/CAMB-77426/2020\|EPI_ISL_439375\|202 | ···················· | ···················· | ····················· | ···················· | ···················· | ···················· |
| B | 4995 | hCoV-19/England/CAMB-772B3/2020\|EPI_ISL_439381\|202 | ···················· | ···················· | ····················· | ···················· | ···················· | ···················· |
| B | 4996 | hCoV-19/England/CAMB-77B7C/2020\|EPI_ISL_439382\|202 | ···················· | ···················· | ····················· | ···················· | ···················· | ···················· |
| B | 4997 | hCoV-19/England/CAMB-7720E/2020\|EPI_ISL_439383\|202 | ···················· | ···················· | ····················· | ···················· | ···················· | ···················· |
| B | 4998 | hCoV-19/England/CAMB-7731A/2020\|EPI_ISL_439384\|202 | ···················· | ···················· | ····················· | ···················· | ···················· | ···················· |
| B | 4999 | hCoV-19/England/CAMB-77286/2020\|EPI_ISL_439385\|202 | ···················· | ···················· | ····················· | ···················· | ···················· | ···················· |
| B | 5000 | hCoV-19/England/CAMB-7780C/2020\|EPI_ISL_439386\|202 | ···················· | ···················· | ····················· | ···················· | ···················· | ···················· |
| B | 5001 | hCoV-19/England/CAMB-77769/2020\|EPI_ISL_439387\|202 | ···················· | ···················· | ····················· | ···················· | ···················· | ···················· |
| B | 5002 | hCoV-19/England/CAMB-77611/2020\|EPI_ISL_439388\|202 | ···················· | ···················· | ····················· | ···················· | ···················· | ···················· |
| B | 5003 | hCoV-19/England/CAMB-76E65/2020\|EPI_ISL_439389\|202 | ···················· | ···················· | ····················· | ···················· | ···················· | ···················· |
| B | 5004 | hCoV-19/England/CAMB-74201/2020\|EPI_ISL_440333\|202 | ···················· | ···················· | ····················· | ···················· | ···················· | ···················· |
| B | 5005 | hCoV-19/England/CAMB-73FFC/2020\|EPI_ISL_440335\|202 | ···················· | ···················· | ····················· | ···················· | ···················· | ···················· |
| B | 5006 | hCoV-19/England/CAMB-741C8/2020\|EPI_ISL_440337\|202 | ···················· | ···················· | ····················· | ···················· | ···················· | ···················· |
| B | 5007 | hCoV-19/England/CAMB-741E6/2020\|EPI_ISL_440338\|202 | ···················· | ···················· | ····················· | ···················· | ···················· | ···················· |
| B | 5008 | hCoV-19/England/CAMB-7408F/2020\|EPI_ISL_440340\|202 | ···················· | ···················· | ····················· | ···················· | ···················· | ···················· |
| B | 5009 | hCoV-19/England/CAMB-73E95/2020\|EPI_ISL_440341\|202 | ···················· | ···················· | ····················· | ···················· | ···················· | ···················· |
| B | 5010 | hCoV-19/England/CAMB-73EA4/2020\|EPI_ISL_440342\|202 | ···················· | ···················· | ····················· | ···················· | ···················· | ···················· |
| B | 5011 | hCoV-19/England/CAMB-73F0B/2020\|EPI_ISL_440343\|202 | ···················· | ···················· | ····················· | ···················· | ···················· | ···················· |
| B | 5012 | hCoV-19/England/CAMB-73E3B/2020\|EPI_ISL_440344\|202 | ···················· | ···················· | ····················· | ···················· | ···················· | ···················· |
| B | 5013 | hCoV-19/England/CAMB-7422F/2020\|EPI_ISL_440345\|202 | ···················· | ···················· | ····················· | ···················· | ···················· | ···················· |
| B | 5014 | hCoV-19/England/CAMB-74104/2020\|EPI_ISL_440347\|202 | ···················· | ···················· | ····················· | ···················· | ···················· | ···················· |
| B | 5015 | hCoV-19/England/CAMB-7425C/2020\|EPI_ISL_440348\|202 | ···················· | ·············A······ | ····················· | ···················· | ···················· | ···················· |
| B | 5016 | hCoV-19/England/CAMB-758A4/2020\|EPI_ISL_440351\|202 | ···················· | ···················· | ····················· | ···················· | ···················· | ···················· |
| B | 5017 | hCoV-19/England/CAMB-74034/2020\|EPI_ISL_440352\|202 | ···················· | ···················· | ····················· | ···················· | ···················· | ···················· |
| B | 5018 | hCoV-19/England/CAMB-73FED/2020\|EPI_ISL_440356\|202 | ···················· | ···················· | ····················· | ···················· | ···················· | ···················· |
| B | 5019 | hCoV-19/England/CAMB-73E77/2020\|EPI_ISL_440357\|202 | ···················· | ···················· | ····················· | ···················· | ···················· | ···················· |
| B | 5020 | hCoV-19/England/CAMB-74007/2020\|EPI_ISL_440358\|202 | ···················· | ···················· | ····················· | ···················· | ···················· | ···················· |
| B | 5021 | hCoV-19/England/CAMB-7426B/2020\|EPI_ISL_440359\|202 | ···················· | ···················· | ····················· | ···················· | ···················· | ···················· |
| B | 5022 | hCoV-19/England/CAMB-73E0E/2020\|EPI_ISL_440365\|202 | ···················· | ···················· | ····················· | ···················· | ···················· | ···················· |
| B | 5023 | hCoV-19/England/CAMB-73EFF/2020\|EPI_ISL_440367\|202 | ···················· | ···················· | ····················· | ···················· | ···················· | ···················· |
| B | 5024 | hCoV-19/England/CAMB-74140/2020\|EPI_ISL_440369\|202 | ···················· | ···················· | ····················· | ···················· | ···················· | ···················· |

| | | | | | | | | |
|---|---|---|---|---|---|---|---|---|
| B | 5025 | hCoV-19/England/CAMB-74025/2020\|EPI_ISL_440373\|202 | .................... | .................... | ..................... | .................... | .................... | .................... |
| B | 5026 | hCoV-19/England/CAMB-751D6/2020\|EPI_ISL_440374\|202 | .................... | .................... | ..................... | .................... | .................... | .................... |
| B | 5027 | hCoV-19/England/CAMB-7524C/2020\|EPI_ISL_440375\|202 | .................... | .................... | ..................... | .................... | .................... | .................... |
| B | 5028 | hCoV-19/England/CAMB-7CD71/2020\|EPI_ISL_441600\|202 | .................... | .................... | ..................... | .................... | .................... | .................... |
| B | 5029 | hCoV-19/England/CAMB-7DF4D/2020\|EPI_ISL_441602\|202 | .................... | .................... | ..................... | .................... | .................... | .................... |
| B | 5030 | hCoV-19/England/CAMB-7CEAB/2020\|EPI_ISL_441603\|202 | .................... | .................... | ..................... | .................... | .................... | .................... |
| B | 5031 | hCoV-19/England/CAMB-7B9E7/2020\|EPI_ISL_441604\|202 | .................... | .................... | ..................... | .................... | .................... | .................... |
| B | 5032 | hCoV-19/England/CAMB-7D0FF/2020\|EPI_ISL_441605\|202 | .................... | .................... | ..................... | .................... | .................... | .................... |
| B | 5033 | hCoV-19/England/CAMB-7D314/2020\|EPI_ISL_441606\|202 | .................... | .................... | ..................... | .................... | .................... | .................... |
| B | 5034 | hCoV-19/England/CAMB-7B941/2020\|EPI_ISL_441607\|202 | .................... | .................... | ..................... | .................... | .................... | .................... |
| B | 5035 | hCoV-19/England/CAMB-7BA6C/2020\|EPI_ISL_441609\|202 | .................... | .................... | ..................... | .................... | .................... | .................... |
| B | 5036 | hCoV-19/England/CAMB-7C4C7/2020\|EPI_ISL_441618\|202 | .................... | .................... | ..................... | .................... | .................... | .................... |
| B | 5037 | hCoV-19/Northern Ireland/NIRE-FA810/2020\|EPI_ISL_4 | .................... | .................... | ..................... | .................... | .................... | .................... |
| B | 5038 | hCoV-19/England/CAMB-7D6DF/2020\|EPI_ISL_441664\|202 | .................... | .................... | ..................... | .................... | .................... | .................... |
| B | 5039 | hCoV-19/England/CAMB-7D860/2020\|EPI_ISL_441665\|202 | .................... | .................... | ..................... | .................... | .................... | .................... |
| B | 5040 | hCoV-19/England/CAMB-7DA3D/2020\|EPI_ISL_441666\|202 | .................... | .................... | ..................... | .................... | .................... | .................... |
| B | 5041 | hCoV-19/England/CAMB-7D46C/2020\|EPI_ISL_441667\|202 | .................... | .................... | ..................... | .................... | .................... | .................... |
| B | 5042 | hCoV-19/England/CAMB-7D903/2020\|EPI_ISL_441668\|202 | .................... | .................... | ..................... | .................... | .................... | .................... |
| B | 5043 | hCoV-19/England/CAMB-7DD16/2020\|EPI_ISL_441669\|202 | .................... | .................... | ..................... | .................... | .................... | .................... |
| B | 5044 | hCoV-19/Northern Ireland/NIRE-FA634/2020\|EPI_ISL_4 | .................... | .................... | ..................... | .................... | .................... | .................... |
| B | 5045 | hCoV-19/England/CAMB-7D8BB/2020\|EPI_ISL_441671\|202 | .................... | .................... | ..................... | .................... | .................... | .................... |
| B | 5046 | hCoV-19/England/CAMB-7D6FD/2020\|EPI_ISL_441672\|202 | .................... | .................... | ..................... | .................... | .................... | .................... |
| B | 5047 | hCoV-19/England/CAMB-7DBD0/2020\|EPI_ISL_441673\|202 | .................... | .................... | ..................... | .................... | .................... | .................... |
| B | 5048 | hCoV-19/England/CAMB-7DCFB/2020\|EPI_ISL_441674\|202 | .................... | .................... | ..................... | .................... | .................... | .................... |
| B | 5049 | hCoV-19/Northern Ireland/NIRE-FA7D7/2020\|EPI_ISL_4 | .................... | .................... | ..................... | .................... | .................... | .................... |
| B | 5050 | hCoV-19/Northern Ireland/NIRE-FA93B/2020\|EPI_ISL_4 | .................... | .................... | ..................... | .................... | .................... | .................... |
| B | 5051 | hCoV-19/England/CAMB-7DA97/2020\|EPI_ISL_441677\|202 | .................... | .................... | ..................... | .................... | .................... | .................... |
| B | 5052 | hCoV-19/England/CAMB-7DBB2/2020\|EPI_ISL_441678\|202 | .................... | .................... | ..................... | .................... | .................... | .................... |
| B | 5053 | hCoV-19/England/CAMB-7DC55/2020\|EPI_ISL_441679\|202 | .................... | .................... | ..................... | .................... | .................... | .................... |
| B | 5054 | hCoV-19/England/CAMB-7D5A5/2020\|EPI_ISL_441680\|202 | .................... | .................... | ..................... | .................... | .................... | .................... |
| B | 5055 | hCoV-19/England/CAMB-7DCBF/2020\|EPI_ISL_441681\|202 | .................... | .................... | ..................... | .................... | .................... | .................... |
| B | 5056 | hCoV-19/England/CAMB-7D47B/2020\|EPI_ISL_441682\|202 | .................... | .................... | ..................... | .................... | .................... | .................... |
| B | 5057 | hCoV-19/England/CAMB-7D98B/2020\|EPI_ISL_441683\|202 | .................... | .................... | ..................... | .................... | .................... | .................... |
| B | 5058 | hCoV-19/England/CAMB-7D824/2020\|EPI_ISL_441684\|202 | .................... | .................... | ..................... | .................... | .................... | .................... |
| B | 5059 | hCoV-19/England/CAMB-7DC37/2020\|EPI_ISL_441686\|202 | .................... | .................... | ..................... | .................... | .................... | .................... |
| B | 5060 | hCoV-19/England/CAMB-7D718/2020\|EPI_ISL_441689\|202 | .................... | .................... | ..................... | .................... | .................... | .................... |
| B | 5061 | hCoV-19/Northern Ireland/NIRE-FA79B/2020\|EPI_ISL_4 | .................... | .................... | ..................... | .................... | .................... | .................... |
| B | 5062 | hCoV-19/Northern Ireland/NIRE-FA77D/2020\|EPI_ISL_4 | .................... | .................... | ..................... | .................... | .................... | .................... |
| B | 5063 | hCoV-19/Northern Ireland/NIRE-FA968/2020\|EPI_ISL_4 | .................... | .................... | ..................... | .................... | .................... | .................... |
| B | 5064 | hCoV-19/Northern Ireland/NIRE-FAA0B/2020\|EPI_ISL_4 | .................... | .................... | ..................... | .................... | .................... | .................... |
| B | 5065 | hCoV-19/England/CAMB-7F2BB/2020\|EPI_ISL_442900\|202 | .................... | .................... | ..................... | .................... | .................... | .................... |
| B | 5066 | hCoV-19/England/CAMB-7F0C0/2020\|EPI_ISL_442901\|202 | .................... | .................... | ..................... | .................... | .................... | .................... |
| B | 5067 | hCoV-19/England/CAMB-8085B/2020\|EPI_ISL_442903\|202 | .................... | .................... | ..................... | .................... | .................... | .................... |
| B | 5068 | hCoV-19/England/CAMB-7F303/2020\|EPI_ISL_442904\|202 | .................... | .................... | ..................... | .................... | .................... | .................... |
| B | 5069 | hCoV-19/England/CAMB-81210/2020\|EPI_ISL_442905\|202 | .................... | .................... | ..................... | .................... | .................... | .................... |
| B | 5070 | hCoV-19/England/CAMB-7F093/2020\|EPI_ISL_442906\|202 | .................... | .................... | ..................... | .................... | .................... | .................... |
| B | 5071 | hCoV-19/England/CAMB-811F5/2020\|EPI_ISL_442907\|202 | .................... | .................... | ..................... | .................... | .................... | .................... |
| B | 5072 | hCoV-19/England/CAMB-7EFE2/2020\|EPI_ISL_442908\|202 | .................... | .A.................. | ..................... | .................... | .................... | .................... |
| B | 5073 | hCoV-19/England/CAMB-7EC54/2020\|EPI_ISL_442909\|202 | .................... | .................... | ..................... | .................... | .................... | .................... |
| B | 5074 | hCoV-19/England/CAMB-7EFB5/2020\|EPI_ISL_442930\|202 | .................... | .................... | ..................... | .................... | .................... | .................... |
| B | 5075 | hCoV-19/England/CAMB-7EB66/2020\|EPI_ISL_442932\|202 | .................... | .................... | ..................... | .................... | .................... | .................... |
| B | 5076 | hCoV-19/England/CAMB-812E3/2020\|EPI_ISL_442934\|202 | .................... | .................... | ..................... | .................... | .................... | .................... |
| B | 5077 | hCoV-19/England/CAMB-7F215/2020\|EPI_ISL_442935\|202 | .................... | .................... | ..................... | .................... | .................... | .................... |
| B | 5078 | hCoV-19/England/CAMB-7F2AC/2020\|EPI_ISL_442936\|202 | .................... | .................... | ..................... | .................... | .................... | .................... |
| B | 5079 | hCoV-19/England/CAMB-80D98/2020\|EPI_ISL_442937\|202 | .................... | .................... | ..................... | .................... | .................... | .................... |
| B | 5080 | hCoV-19/England/CAMB-7EC72/2020\|EPI_ISL_442938\|202 | .................... | .................... | ..................... | .................... | .................... | .................... |
| B | 5081 | hCoV-19/England/CAMB-8083D/2020\|EPI_ISL_442939\|202 | .................... | .................... | ..................... | .................... | .................... | .................... |
| B | 5082 | hCoV-19/England/CAMB-7EBA2/2020\|EPI_ISL_442994\|202 | .................... | .................... | ..................... | .................... | .................... | .................... |
| B | 5083 | hCoV-19/England/CAMB-7F594/2020\|EPI_ISL_442995\|202 | .................... | .................... | ..................... | .................... | .................... | .................... |
| B | 5084 | hCoV-19/England/CAMB-80DD4/2020\|EPI_ISL_442996\|202 | .................... | .................... | ..................... | .................... | .................... | .................... |
| B | 5085 | hCoV-19/England/CAMB-7EDE8/2020\|EPI_ISL_442997\|202 | .................... | .................... | ..................... | .................... | .................... | .................... |
| B | 5086 | hCoV-19/England/CAMB-7EB1B/2020\|EPI_ISL_442998\|202 | .................... | .................... | ..................... | .................... | .................... | .................... |
| B | 5087 | hCoV-19/England/CAMB-7EC45/2020\|EPI_ISL_442999\|202 | .................... | .................... | ..................... | .................... | .................... | .................... |
| B | 5088 | hCoV-19/USA/LA-SR0295/2020\|EPI_ISL_445150\|2020-04- | .................... | .................... | ..................... | .................... | .................... | .................... |
| B | 5089 | hCoV-19/USA/LA-SR0297/2020\|EPI_ISL_445152\|2020-04- | .................... | .................... | ..................... | .................... | .................... | .................... |
| B | 5090 | hCoV-19/USA/LA-SR0298/2020\|EPI_ISL_445153\|2020-04- | .................... | .................... | ..................... | .................... | .................... | .................... |
| B | 5091 | hCoV-19/USA/LA-SR0299/2020\|EPI_ISL_445154\|2020-04- | .................... | .................... | ..................... | .................... | .................... | .................... |
| B | 5092 | hCoV-19/USA/LA-SR0300/2020\|EPI_ISL_445155\|2020-04- | .................... | .................... | ..................... | .................... | .................... | .................... |
| B | 5093 | hCoV-19/USA/LA-SR0301/2020\|EPI_ISL_445156\|2020-04- | .................... | .................... | ..................... | .................... | .................... | .................... |
| B | 5094 | hCoV-19/USA/LA-SR0302/2020\|EPI_ISL_445157\|2020-04- | .................... | .................... | ..................... | .................... | .................... | .................... |
| B | 5095 | hCoV-19/USA/LA-SR0304/2020\|EPI_ISL_445159\|2020-04- | .................... | .................... | ..................... | .................... | .................... | .................... |
| B | 5096 | hCoV-19/USA/LA-SR0306/2020\|EPI_ISL_445161\|2020-04- | .................... | .................... | ..................... | .................... | .................... | .................... |
| B | 5097 | hCoV-19/USA/LA-SR0307/2020\|EPI_ISL_445162\|2020-04- | .................... | .................... | ..................... | .................... | .................... | .................... |
| B | 5098 | hCoV-19/USA/CA-SR0310/2020\|EPI_ISL_445164\|2020-03- | .................... | .................... | ..................... | .................... | .................... | .................... |
| B | 5099 | hCoV-19/USA/CA-SR0312/2020\|EPI_ISL_445166\|2020-03- | .................... | .................... | ..................... | .................... | .................... | .................... |
| B | 5100 | hCoV-19/USA/CA-SR0313/2020\|EPI_ISL_445167\|2020-03- | .................... | .................... | ..................... | .................... | .................... | .................... |
| B | 5101 | hCoV-19/USA/CA-CZB-1121/2020\|EPI_ISL_445169\|2020-0 | .................... | .................... | ..................... | .................... | .................... | .................... |
| B | 5102 | hCoV-19/USA/CA-CZB-1123/2020\|EPI_ISL_445171\|2020-0 | .................... | .................... | ..................... | .................... | .................... | .................... |

| | | | | | | | | |
|---|---|---|---|---|---|---|---|---|
| B | 5103 hCoV-19/USA/CA-CZB-1124/2020\|EPI_ISL_445172\|2020-0 | ···················· | ···················· | ···················· | ···················· | ···················· | ···················· | |
| B | 5104 hCoV-19/USA/CA-CZB-1125/2020\|EPI_ISL_445173\|2020-0 | ···················· | ···················· | ···················· | ···················· | ···················· | ···················· | |
| B | 5105 hCoV-19/USA/CA-CZB-1127/2020\|EPI_ISL_445175\|2020-0 | ···················· | ···················· | ···················· | ···················· | ···················· | ···················· | |
| B | 5106 hCoV-19/USA/CA-CZB-1128/2020\|EPI_ISL_445176\|2020-0 | ···················· | ···················· | ···················· | ···················· | ···················· | ···················· | |
| B | 5107 hCoV-19/USA/CA-CZB-1129/2020\|EPI_ISL_445177\|2020-0 | ···················· | ···················· | ···················· | ···················· | ···················· | ···················· | |
| B | 5108 hCoV-19/USA/CA-CZB-1131/2020\|EPI_ISL_445179\|2020-0 | ···················· | ···················· | ···················· | ···················· | ···················· | ···················· | |
| B | 5109 hCoV-19/USA/CA-CZB-1132/2020\|EPI_ISL_445180\|2020-0 | ···················· | ···················· | ···················· | ···················· | ···················· | ···················· | |
| B | 5110 hCoV-19/USA/CA-CZB-1133/2020\|EPI_ISL_445181\|2020-0 | ···················· | ···················· | ···················· | ···················· | ···················· | ···················· | |
| B | 5111 hCoV-19/USA/CA-CZB-1134/2020\|EPI_ISL_445182\|2020-0 | ···················· | ···················· | ···················· | ···················· | ···················· | ···················· | |
| B | 5112 hCoV-19/Japan/Hu_Kng_19-865/2020\|EPI_ISL_445183\|20 | ···················· | ···················· | ···················· | ···················· | ···················· | ···················· | |
| B | 5113 hCoV-19/Wales/PHWC-3177A/2020\|EPI_ISL_446483\|2020- | ···················· | ···················· | ···················· | ···················· | ···················· | ···················· | |
| B | 5114 hCoV-19/Wales/PHWC-31789/2020\|EPI_ISL_446484\|2020- | ···················· | ···A················ | ···················· | ···················· | ···················· | ···················· | |
| B | 5115 hCoV-19/Wales/PHWC-317A7/2020\|EPI_ISL_446486\|2020- | ···················· | ···················· | ···················· | ···················· | ···················· | ···················· | |
| B | 5116 hCoV-19/Wales/PHWC-3181D/2020\|EPI_ISL_446493\|2020- | ···················· | ···················· | ···················· | ···················· | ···················· | ···················· | |
| B | 5117 hCoV-19/Wales/PHWC-3182C/2020\|EPI_ISL_446494\|2020- | ···················· | ···A················ | ···················· | ···················· | ···················· | ···················· | |
| B | 5118 hCoV-19/Wales/PHWC-3184A/2020\|EPI_ISL_446496\|2020- | ···················· | ···················· | ···················· | ···················· | ···················· | ···················· | |
| B | 5119 hCoV-19/Wales/PHWC-31859/2020\|EPI_ISL_446497\|2020- | ···················· | ···A················ | ···················· | ···················· | ···················· | ···················· | |
| B | 5120 hCoV-19/Wales/PHWC-31895/2020\|EPI_ISL_446498\|2020- | ···················· | ···················· | ···················· | ···················· | ···················· | ···················· | |
| B | 5121 hCoV-19/USA/WA-UW-5911/2020\|EPI_ISL_430930\|2020-04 | ···················· | ···················· | ···················· | ···················· | ···················· | ···················· | |
| B | 5122 hCoV-19/USA/WA-UW-5932/2020\|EPI_ISL_430931\|2020-04 | ···················· | ···················· | ···················· | ···················· | ···················· | ···················· | |
| B | 5123 hCoV-19/USA/WA-UW-5947/2020\|EPI_ISL_430932\|2020-04 | ···················· | ···················· | ···················· | ···················· | ···················· | ···················· | |
| B | 5124 hCoV-19/USA/WA-UW-6162/2020\|EPI_ISL_430933\|2020-04 | ···················· | ···················· | ···················· | ···················· | ···················· | ···················· | |
| B | 5125 hCoV-19/USA/WA-UW-6164/2020\|EPI_ISL_430934\|2020-04 | ···················· | ···················· | ···················· | ···················· | ···················· | ···················· | |
| B | 5126 hCoV-19/USA/WA-UW-6173/2020\|EPI_ISL_430936\|2020-04 | ···················· | ···················· | ···················· | ···················· | ···················· | ···················· | |
| B | 5127 hCoV-19/USA/WA-UW-6178/2020\|EPI_ISL_430937\|2020-04 | ···················· | ···················· | ···················· | ···················· | ···················· | ···················· | |
| B | 5128 hCoV-19/USA/WA-UW-4226/2020\|EPI_ISL_430938\|2020-03 | ···················· | ···················· | ···················· | ···················· | ···················· | ···················· | |
| B | 5129 hCoV-19/USA/WA-UW-4243/2020\|EPI_ISL_430939\|2020-04 | ···················· | ···················· | ···················· | ···················· | ···················· | ···················· | |
| B | 5130 hCoV-19/USA/ID-UW-4255/2020\|EPI_ISL_430940\|2020-03 | ···················· | ···················· | ···················· | ···················· | ···················· | ···················· | |
| B | 5131 hCoV-19/USA/WA-UW-4279/2020\|EPI_ISL_430941\|2020-03 | ···················· | ···················· | ···················· | ···················· | ···················· | ···················· | |
| B | 5132 hCoV-19/USA/ID-UW-4307/2020\|EPI_ISL_430942\|2020-03 | ···················· | ···················· | ···················· | ···················· | ···················· | ···················· | |
| B | 5133 hCoV-19/USA/WA-UW-4338/2020\|EPI_ISL_430943\|2020-03 | ···················· | ···················· | ···················· | ···················· | ···················· | ···················· | |
| B | 5134 hCoV-19/USA/CT-UW-4345/2020\|EPI_ISL_430945\|2020-03 | ···················· | ···················· | ···················· | ···················· | ···················· | ···················· | |
| B | 5135 hCoV-19/USA/CT-UW-4348/2020\|EPI_ISL_430946\|2020-03 | ···················· | ···················· | ···················· | ···················· | ···················· | ···················· | |
| B | 5136 hCoV-19/USA/ID-UW-4354/2020\|EPI_ISL_430947\|2020-03 | ···················· | ···················· | ···················· | ···················· | ···················· | ···················· | |
| B | 5137 hCoV-19/USA/ID-UW-4383/2020\|EPI_ISL_430948\|2020-03 | ···················· | ···················· | ···················· | ···················· | ···················· | ···················· | |
| B | 5138 hCoV-19/USA/ID-UW-4402/2020\|EPI_ISL_430949\|2020-03 | ···················· | ···················· | ···················· | ···················· | ···················· | ···················· | |
| B | 5139 hCoV-19/USA/ID-UW-6349/2020\|EPI_ISL_430970\|2020-04 | ···················· | ···················· | ···················· | ···················· | ···················· | ···················· | |
| B | 5143 hCoV-19/Scotland/EDB1647/2020\|EPI_ISL_433104\|2020- | ···················· | ···················· | ···················· | ···················· | ···················· | ···················· | |
| B | 5146 hCoV-19/Scotland/EDB1655/2020\|EPI_ISL_433107\|2020- | ···················· | ···················· | ···················· | ···················· | ···················· | ···················· | |
| B | 5147 hCoV-19/Scotland/EDB1656/2020\|EPI_ISL_433108\|2020- | ···················· | ···········C········ | ···················· | ···················· | ···················· | ···················· | |
| B | 5149 hCoV-19/Scotland/EDB1663/2020\|EPI_ISL_433110\|2020- | ···················· | ···················· | ···················· | ···················· | ···················· | ···················· | |
| B | 5151 hCoV-19/Scotland/EDB1671/2020\|EPI_ISL_433112\|2020- | ···················· | ···················· | ···················· | ···················· | ···················· | ···················· | |
| B | 5152 hCoV-19/Scotland/EDB1674/2020\|EPI_ISL_433113\|2020- | ···················· | ···················A | ···················· | ···················· | ···················· | ···················· | 1 |
| B | 5153 hCoV-19/Scotland/EDB1677/2020\|EPI_ISL_433114\|2020- | ···················· | ···················· | ···················· | ···················· | ···················· | ···················· | |
| B | 5154 hCoV-19/Scotland/EDB1679/2020\|EPI_ISL_433115\|2020- | ···················· | ·A·················· | ···················· | ···················· | ···················· | ···················· | |
| B | 5155 hCoV-19/Scotland/EDB1681/2020\|EPI_ISL_433116\|2020- | ···················· | ·A·················· | ···················· | ···················· | ···················· | ···················· | |
| B | 5156 hCoV-19/Scotland/EDB1683/2020\|EPI_ISL_433117\|2020- | ···················· | ···················· | ···················· | ···················· | ···················· | ···················· | |
| B | 5157 hCoV-19/Scotland/EDB1693/2020\|EPI_ISL_433118\|2020- | ···················· | ···················· | ···················· | ···················· | ···················· | ···················· | |
| B | 5158 hCoV-19/Scotland/EDB1694/2020\|EPI_ISL_433119\|2020- | ···················· | ···················· | ···················· | ···················· | ···················· | ···················· | |
| B | 5159 hCoV-19/Scotland/EDB1697/2020\|EPI_ISL_433120\|2020- | ···················· | ·A·················· | ···················· | ···················· | ···················· | ···················· | |
| B | 5160 hCoV-19/Scotland/EDB1700/2020\|EPI_ISL_433121\|2020- | ···················· | ···················· | ···················· | ···················· | ···················· | ···················· | |
| B | 5161 hCoV-19/Scotland/EDB1701/2020\|EPI_ISL_433122\|2020- | ···················· | ···················· | ···················· | ···················· | ···················· | ···················· | |
| B | 5162 hCoV-19/Scotland/EDB1704/2020\|EPI_ISL_433123\|2020- | ···················· | ···················· | ···················· | ···················· | ···················· | ···················· | |
| B | 5163 hCoV-19/Scotland/EDB1706/2020\|EPI_ISL_433124\|2020- | ···················· | ···················· | ···················· | ···················· | ···················· | ···················· | |
| B | 5164 hCoV-19/Scotland/EDB1707/2020\|EPI_ISL_433125\|2020- | ···················· | ·A·················· | ···················· | ···················· | ···················· | ···················· | |
| B | 5165 hCoV-19/Scotland/EDB1711/2020\|EPI_ISL_433126\|2020- | ···················· | ···················A | ···················· | ···················· | ···················· | ···················· | 1 |
| B | 5166 hCoV-19/Scotland/EDB1717/2020\|EPI_ISL_433127\|2020- | ···················· | ···················· | ···················· | ···················· | ···················· | ···················· | |
| B | 5167 hCoV-19/Scotland/EDB1725/2020\|EPI_ISL_433129\|2020- | ···················· | ·A·················· | ···················· | ···················· | ···················· | ···················· | |
| B | 5168 hCoV-19/Scotland/EDB1730/2020\|EPI_ISL_433130\|2020- | ···················· | ···················· | ···················· | ···················· | ···················· | ···················· | |
| B | 5169 hCoV-19/Scotland/EDB1732/2020\|EPI_ISL_433131\|2020- | ···················· | ···················· | ···················· | ···················· | ···················· | ···················· | |
| B | 5170 hCoV-19/Scotland/EDB1738/2020\|EPI_ISL_433132\|2020- | ···················· | ···················· | ···················· | ···················· | ···················· | ···················· | |
| B | 5171 hCoV-19/Scotland/EDB1740/2020\|EPI_ISL_433133\|2020- | ···················· | ·A·················· | ···················· | ···················· | ···················· | ···················· | |
| B | 5172 hCoV-19/Scotland/EDB1811/2020\|EPI_ISL_433135\|2020- | ···················· | ···················· | ···················· | ···················· | ···················· | ···················· | |
| B | 5173 hCoV-19/Scotland/EDB1812/2020\|EPI_ISL_433136\|2020- | ···················· | ···················· | ···················· | ···················· | ···················· | ···················· | |
| B | 5174 hCoV-19/Scotland/EDB1813/2020\|EPI_ISL_433137\|2020- | ···················· | ···················· | ···················· | ···················· | ···················· | ···················· | |
| B | 5175 hCoV-19/Scotland/EDB1816/2020\|EPI_ISL_433139\|2020- | ···················· | ···················A | ···················· | ···················· | ···················· | ···················· | 1 |
| B | 5176 hCoV-19/Scotland/EDB1817/2020\|EPI_ISL_433140\|2020- | ···················· | ···················· | ···················· | ···················· | ···················· | ···················· | |
| B | 5177 hCoV-19/Scotland/EDB1819/2020\|EPI_ISL_433141\|2020- | ···················· | ···················· | ···················· | ···················· | ···················· | ···················· | |
| B | 5178 hCoV-19/Scotland/EDB1820/2020\|EPI_ISL_433142\|2020- | ···················· | ···················· | ···················· | ···················· | ···················· | ···················· | |
| B | 5179 hCoV-19/Scotland/EDB1821/2020\|EPI_ISL_433143\|2020- | ···················· | ·A·················· | ···················· | ···················· | ···················· | ···················· | |
| B | 5180 hCoV-19/Scotland/EDB1825/2020\|EPI_ISL_433145\|2020- | ···················· | ···················· | ···················· | ···················· | ···················· | ···················· | |
| B | 5181 hCoV-19/Scotland/EDB1827/2020\|EPI_ISL_433147\|2020- | ···················· | ·A·················· | ···················· | ···················· | ···················· | ···················· | |
| B | 5182 hCoV-19/Scotland/EDB1828/2020\|EPI_ISL_433148\|2020- | ···················· | ·A·················· | ···················· | ···················· | ···················· | ···················· | |
| B | 5183 hCoV-19/Scotland/EDB1829/2020\|EPI_ISL_433149\|2020- | ···················· | ·A·················· | ···················· | ···················· | ···················· | ···················· | |
| B | 5184 hCoV-19/Scotland/EDB1830/2020\|EPI_ISL_433150\|2020- | ···················· | ·A·················· | ···················· | ···················· | ···················· | ···················· | |
| B | 5185 hCoV-19/Scotland/EDB1841/2020\|EPI_ISL_433153\|2020- | ···················· | ·A·················· | ···················· | ···················· | ···················· | ···················· | |
| B | 5186 hCoV-19/Scotland/EDB1842/2020\|EPI_ISL_433154\|2020- | ···················· | ·A·················· | ···················· | ···················· | ···················· | ···················· | |
| B | 5187 hCoV-19/Scotland/EDB1848/2020\|EPI_ISL_433155\|2020- | ···················· | ·A·················· | ···················· | ···················· | ···················· | ···················· | |

| | | | | | | | |
|---|---|---|---|---|---|---|---|
| B | 5188 hCoV-19/Scotland/EDB1897/2020\|EPI_ISL_433156\|2020- | ···················· | ···················· | ···················· | ···················· | ···················· | ···················· |
| B | 5189 hCoV-19/Scotland/EDB2011/2020\|EPI_ISL_433162\|2020- | ···················· | ·A·················· | ···················· | ···················· | ···················· | ···················· |
| B | 5190 hCoV-19/Scotland/EDB2048/2020\|EPI_ISL_433164\|2020- | ···················· | ·A·················· | ···················· | ···················· | ···················· | ···················· |
| B | 5191 hCoV-19/Scotland/EDB2049/2020\|EPI_ISL_433165\|2020- | ···················· | ···················· | ···················· | ···················· | ···················· | ···················· |
| B | 5192 hCoV-19/Scotland/EDB2050/2020\|EPI_ISL_433166\|2020- | ·T·················· | ···················· | ···················· | ···················· | ···················· | ···················· |
| B | 5193 hCoV-19/Scotland/EDB2051/2020\|EPI_ISL_433167\|2020- | ···················· | ···················· | ···················· | ···················· | ···················· | ···················· |
| B | 5194 hCoV-19/Scotland/EDB2052/2020\|EPI_ISL_433168\|2020- | ···················· | ···················· | ···················· | ···················· | ···················· | ···················· |
| B | 5195 hCoV-19/Scotland/EDB2057/2020\|EPI_ISL_433169\|2020- | ···················· | ·A·················· | ···················· | ···················· | ···················· | ···················· |
| B | 5196 hCoV-19/Scotland/CVR512/2020\|EPI_ISL_433308\|2020-0 | ···················· | ···················· | ···················· | ···················· | ···················· | ···················· |
| B | 5197 hCoV-19/Greece/133/2020\|EPI_ISL_434455\|2020-03-09 | ···················· | ···················· | ···················· | ···················· | ···················· | ···················· |
| B | 5198 hCoV-19/Greece/48/2020\|EPI_ISL_434456\|2020-03-22 | ···················· | ···················· | ···················· | ···················· | ···················· | ···················· |
| B | 5199 hCoV-19/Greece/36/2020\|EPI_ISL_434457\|2020-03-28 | ···················· | ···················· | ···················· | ···················· | ···················· | ···················· |
| B | 5200 hCoV-19/Greece/37/2020\|EPI_ISL_434458\|2020-03-20 | ···················· | ···················· | ···················· | ···················· | ···················· | ···················· |
| B | 5201 hCoV-19/Greece/35/2020\|EPI_ISL_434459\|2020-03-29 | ···················· | ···················· | ···················· | ···················· | ···················· | ···················· |
| B | 5202 hCoV-19/Greece/38/2020\|EPI_ISL_434460\|2020-03-23 | ···················· | ···················· | ···················· | ···················· | ···················· | ···················· |
| B | 5203 hCoV-19/Greece/39/2020\|EPI_ISL_434461\|2020-03-29 | ···················· | ···················· | ···················· | ···················· | ···················· | ···················· |
| B | 5204 hCoV-19/Greece/47/2020\|EPI_ISL_434462\|2020-03-23 | ···················· | ···················· | ···················· | ···················· | ···················· | ···················· |
| B | 5205 hCoV-19/Greece/46/2020\|EPI_ISL_434463\|2020-03-23 | ···················· | ···················· | ···················· | ···················· | ···················· | ···················· |
| B | 5206 hCoV-19/Greece/145/2020\|EPI_ISL_434464\|2020-03-16 | ···················· | ···················· | ···················· | ···················· | ···················· | ···················· |
| B | 5207 hCoV-19/Greece/44/2020\|EPI_ISL_434465\|2020-03-23 | ···················· | ···················· | ···················· | ···················· | ···················· | ···················· |
| B | 5208 hCoV-19/Greece/53/2020\|EPI_ISL_434466\|2020-04-04 | ···················· | ···················· | ···················· | ···················· | ···················· | ···················· |
| B | 5209 hCoV-19/Greece/141/2020\|EPI_ISL_434467\|2020-03-12 | ···················· | ···················· | ···················· | ···················· | ···················· | ···················· |
| B | 5210 hCoV-19/Greece/51/2020\|EPI_ISL_434468\|2020-03-26 | ···················· | ···················· | ···················· | ···················· | ···················· | ···················· |
| B | 5211 hCoV-19/Greece/154/2020\|EPI_ISL_434469\|2020-03-20 | ···················· | ···················· | ···················· | ···················· | ···················· | ···················· |
| B | 5212 hCoV-19/Greece/155/2020\|EPI_ISL_434470\|2020-03-19 | ···················· | ···················· | ···················· | ···················· | ···················· | ···················· |
| B | 5213 hCoV-19/Greece/149/2020\|EPI_ISL_434471\|2020-03-11 | ···················· | ···················· | ···················· | ···················· | ···················· | ···················· |
| B | 5214 hCoV-19/Greece/139/2020\|EPI_ISL_434472\|2020-03-12 | ···················· | ···················· | ···················· | ···················· | ···················· | ···················· |
| B | 5215 hCoV-19/Greece/136/2020\|EPI_ISL_434473\|2020-03-11 | ···················· | ···················· | ···················· | ···················· | ···················· | ···················· |
| B | 5216 hCoV-19/Greece/137/2020\|EPI_ISL_434474\|2020-03-11 | ···················· | ···················· | ···················· | ···················· | ···················· | ···················· |
| B | 5217 hCoV-19/Greece/134/2020\|EPI_ISL_434475\|2020-03-10 | ···················· | ·············A······ | ···················· | ···················· | ···················· | ···················· |
| B | 5218 hCoV-19/Greece/135/2020\|EPI_ISL_434476\|2020-03-10 | ···················· | ···················· | ···················· | ···················· | ···················· | ···················· |
| B | 5219 hCoV-19/Greece/143/2020\|EPI_ISL_434477\|2020-03-16 | ···················· | ···················· | ···················· | ···················· | ···················· | ···················· |
| B | 5220 hCoV-19/Greece/142/2020\|EPI_ISL_434478\|2020-03-14 | ···················· | ···················· | ···················· | ···················· | ···················· | ···················· |
| B | 5221 hCoV-19/Greece/138/2020\|EPI_ISL_434479\|2020-03-11 | ···················· | ···················· | ···················· | ···················· | ···················· | ···················· |
| B | 5222 hCoV-19/Greece/140/2020\|EPI_ISL_434480\|2020-03-12 | ···················· | ···················· | ···················· | ···················· | ···················· | ···················· |
| B | 5223 hCoV-19/Greece/146/2020\|EPI_ISL_434481\|2020-03-19 | ···················· | ···················· | ···················· | ···················· | ···················· | ···················· |
| B | 5224 hCoV-19/Greece/156/2020\|EPI_ISL_434482\|2020-03-16 | ···················· | ···················· | ···················· | ···················· | ···················· | ···················· |
| B | 5225 hCoV-19/Greece/144/2020\|EPI_ISL_434483\|2020-03-16 | ···················· | ···················· | ···················· | ···················· | ···················· | ···················· |
| B | 5226 hCoV-19/Greece/148/2020\|EPI_ISL_434484\|2020-03-09 | ···················· | ···················· | ···················· | ···················· | ···················· | ···················· |
| B | 5227 hCoV-19/Greece/150/2020\|EPI_ISL_434485\|2020-03-12 | ···················· | ···················· | ···················· | ···················· | ···················· | ···················· |
| B | 5228 hCoV-19/Greece/151/2020\|EPI_ISL_434486\|2020-03-13 | ···················· | ···················· | ···················· | ···················· | ···················· | ···················· |
| B | 5229 hCoV-19/Luxembourg/LNS0972189/2020\|EPI_ISL_434487\| | ···················· | ···················· | ···················· | ···················· | ···················· | ···················· |
| B | 5230 hCoV-19/Luxembourg/LNS1673015/2020\|EPI_ISL_434488\| | ···················· | ···················· | ···················· | ···················· | ···················· | ···················· |
| B | 5231 hCoV-19/Luxembourg/LNS8742871/2020\|EPI_ISL_434489\| | ···················· | ···················· | ···················· | ···················· | ···················· | ···················· |
| B | 5232 hCoV-19/Luxembourg/LNS8258882/2020\|EPI_ISL_434491\| | ···················· | ···················· | ···················· | ···················· | ···················· | ···················· |
| B | 5233 hCoV-19/Luxembourg/LNS8655659/2020\|EPI_ISL_434492\| | ···················· | ···················· | ···················· | ···················· | ···················· | ···················· |
| B | 5234 hCoV-19/Luxembourg/LNS1368952/2020\|EPI_ISL_434493\| | ···················· | ···················· | ···················· | ···················· | ···················· | ···················· |
| B | 5235 hCoV-19/Luxembourg/LNS0188413/2020\|EPI_ISL_434495\| | ···················· | ···················· | ···················· | ···················· | ···················· | ···················· |
| B | 5236 hCoV-19/Luxembourg/LNS0361493/2020\|EPI_ISL_434496\| | ···················· | ···················· | ···················· | ···················· | ···················· | ···················· |
| B | 5237 hCoV-19/Luxembourg/LNS2209136/2020\|EPI_ISL_434497\| | ···················· | ···················· | ···················· | ···················· | ···················· | ···················· |
| B | 5238 hCoV-19/Luxembourg/LNS4757989/2020\|EPI_ISL_434498\| | ···················· | ···················· | ···················· | ···················· | ···················· | ···················· |
| B | 5239 hCoV-19/USA/VA-DCLS-0108/2020\|EPI_ISL_434600\|2020- | ···················· | ···················· | ···················· | ···················· | ···················· | ···················· |
| B | 5240 hCoV-19/USA/VA-DCLS-0109/2020\|EPI_ISL_434601\|2020- | ···················· | ···················· | ···················· | ···················· | ···················· | ···················· |
| B | 5241 hCoV-19/USA/VA-DCLS-0110/2020\|EPI_ISL_434602\|2020- | ···················· | ···················· | ···················· | ···················· | ···················· | ···················· |
| B | 5242 hCoV-19/USA/VA-DCLS-0111/2020\|EPI_ISL_434603\|2020- | ···················· | ···················· | ···················· | ···················· | ···················· | ···················· |
| B | 5243 hCoV-19/USA/VA-DCLS-0112/2020\|EPI_ISL_434604\|2020- | ···················· | ···················· | ···················· | ···················· | ···················· | ···················· |
| B | 5244 hCoV-19/USA/VA-DCLS-0114/2020\|EPI_ISL_434605\|2020- | ···················· | ···················· | ···················· | ···················· | ···················· | ···················· |
| B | 5245 hCoV-19/USA/WI-UW-261/2020\|EPI_ISL_434607\|2020-03- | ···················· | ···················· | ···················· | ···················· | ···················· | ···················· |
| B | 5246 hCoV-19/USA/WI-UW-262/2020\|EPI_ISL_434608\|2020-04- | ···················· | ···················· | ···················· | ···················· | ···················· | ···················· |
| B | 5247 hCoV-19/USA/WI-UW-263/2020\|EPI_ISL_434609\|2020-04- | ···················· | ···················· | ···················· | ···················· | ···················· | ···················· |
| B | 5248 hCoV-19/USA/WI-UW-264/2020\|EPI_ISL_434610\|2020-04- | ···················· | ···················· | ···················· | ···················· | ···················· | ···················· |
| B | 5249 hCoV-19/USA/WI-UW-265/2020\|EPI_ISL_434611\|2020-04- | ···················· | ···················· | ···················· | ···················· | ···················· | ···················· |
| B | 5250 hCoV-19/USA/DC-HP00043/2020\|EPI_ISL_434639\|2020-03 | ···················· | ···················· | ···················· | ···················· | ···················· | ···················· |
| B | 5251 hCoV-19/USA/WA-S627/2020\|EPI_ISL_438180\|2020-04-18 | ···················· | ···················· | ···················· | ···················· | ···················· | ···················· |
| B | 5252 hCoV-19/USA/WA-S628/2020\|EPI_ISL_438181\|2020-04-18 | ···················· | ···················· | ···················· | ···················· | ···················· | ···················· |
| B | 5253 hCoV-19/USA/WA-S629/2020\|EPI_ISL_438182\|2020-04-18 | ···················· | ···················· | ···················· | ···················· | ···················· | ···················· |
| B | 5254 hCoV-19/USA/WA-S630/2020\|EPI_ISL_438183\|2020-04-18 | ···················· | ···················· | ···················· | ···················· | ···················· | ···················· |
| B | 5255 hCoV-19/USA/WA-S631/2020\|EPI_ISL_438184\|2020-04-17 | ···················· | ···················· | ···················· | ···················· | ···················· | ···················· |
| B | 5256 hCoV-19/USA/WA-S632/2020\|EPI_ISL_438185\|2020-04-17 | ···················· | ···················· | ···················· | ···················· | ···················· | ···················· |
| B | 5257 hCoV-19/USA/WA-S633/2020\|EPI_ISL_438186\|2020-04-17 | ···················· | ···················· | ···················· | ···················· | ···················· | ···················· |
| B | 5258 hCoV-19/USA/WA-S634/2020\|EPI_ISL_438187\|2020-04-17 | ···················· | ···················· | ···················· | ···················· | ···················· | ···················· |
| B | 5259 hCoV-19/USA/WA-S635/2020\|EPI_ISL_438188\|2020-04-17 | ···················· | ···················· | ···················· | ···················· | ···················· | ···················· |
| B | 5260 hCoV-19/USA/WA-S636/2020\|EPI_ISL_438189\|2020-04-17 | ···················· | ···················· | ···················· | ···················· | ···················· | ···················· |
| B | 5261 hCoV-19/USA/WA-S637/2020\|EPI_ISL_438190\|2020-04-17 | ···················· | ···················· | ···················· | ···················· | ···················· | ···················· |
| B | 5262 hCoV-19/USA/WA-S638/2020\|EPI_ISL_438191\|2020-04-17 | ···················· | ···················· | ···················· | ···················· | ···················· | ···················· |
| B | 5263 hCoV-19/USA/WA-S639/2020\|EPI_ISL_438192\|2020-04-16 | ···················· | ···················· | ···················· | ···················· | ···················· | ···················· |
| B | 5264 hCoV-19/USA/WA-S640/2020\|EPI_ISL_438193\|2020-04-18 | ···················· | ···················· | ···················· | ···················· | ···················· | ···················· |
| B | 5265 hCoV-19/USA/WA-S641/2020\|EPI_ISL_438194\|2020-04-18 | ···················· | ···················· | ···················· | ···················· | ···················· | ···················· |

| | | | | | | | | |
|---|---|---|---|---|---|---|---|---|
| B | 5266 | hCoV-19/USA/WA-S642/2020\|EPI_ISL_438195\|2020-04-19 | ···················· | ···················· | ···················· | ···················· | ···················· | ···················· |
| B | 5267 | hCoV-19/USA/WA-S643/2020\|EPI_ISL_438196\|2020-04-17 | ···················· | ···················· | ···················· | ···················· | ···················· | ···················· |
| B | 5268 | hCoV-19/USA/WA-S644/2020\|EPI_ISL_438197\|2020-04-17 | ···················· | ···················· | ···················· | ···················· | ···················· | ···················· |
| B | 5269 | hCoV-19/USA/WA-S645/2020\|EPI_ISL_438198\|2020-04-17 | ···················· | ···················· | ···················· | ···················· | ···················· | ···················· |
| B | 5270 | hCoV-19/USA/WA-S646/2020\|EPI_ISL_438199\|2020-04-19 | ···················· | ···················· | ···················· | ···················· | ···················· | ···················· |
| B | 5272 | hCoV-19/Scotland/EDB3631/2020\|EPI_ISL_439301\|2020- | ···················· | ···················· | ···················· | ···················· | ···················· | ···················· |
| B | 5274 | hCoV-19/Scotland/EDB3645/2020\|EPI_ISL_439303\|2020- | ···················· | ·A·················· | ···················· | ···················· | ···················· | ···················· |
| B | 5275 | hCoV-19/Scotland/EDB3652/2020\|EPI_ISL_439304\|2020- | ···················· | ···················· | ···················· | ···················· | ···················· | ···················· |
| B | 5276 | hCoV-19/Scotland/EDB3653/2020\|EPI_ISL_439305\|2020- | ···················· | ·A·················· | ···················· | ···················· | ···················· | ···················· |
| B | 5277 | hCoV-19/Scotland/EDB3655/2020\|EPI_ISL_439306\|2020- | ···················· | ···················· | ···················· | ···················· | ···················· | ···················· |
| B | 5278 | hCoV-19/Scotland/EDB3656/2020\|EPI_ISL_439307\|2020- | ···················· | ·A·················· | ···················· | ···················· | ···················· | ···················· |
| B | 5279 | hCoV-19/England/CAMB-7DFC5/2020\|EPI_ISL_441610\|202 | ···················· | ···················· | ···················· | ···················· | ···················· | ···················· |
| B | 5280 | hCoV-19/England/CAMB-7CCED/2020\|EPI_ISL_441611\|202 | ···················· | ···················· | ···················· | ···················· | ···················· | ···················· |
| B | 5281 | hCoV-19/England/CAMB-7B9AB/2020\|EPI_ISL_441612\|202 | ···················· | ···················· | ···················· | ···················· | ···················· | ···················· |
| B | 5282 | hCoV-19/England/CAMB-7C852/2020\|EPI_ISL_441613\|202 | ···················· | ···················· | ···················· | ···················· | ···················· | ···················· |
| B | 5283 | hCoV-19/England/CAMB-7D3C9/2020\|EPI_ISL_441614\|202 | ···················· | ···················· | ···················· | ···················· | ···················· | ···················· |
| B | 5284 | hCoV-19/England/CAMB-7D51E/2020\|EPI_ISL_441615\|202 | ···················· | ···················· | ···················· | ···················· | ···················· | ···················· |
| B | 5285 | hCoV-19/England/CAMB-7C63A/2020\|EPI_ISL_441616\|202 | ···················· | ···················· | ···················· | ···················· | ···················· | ···················· |
| B | 5286 | hCoV-19/England/CAMB-7DF6B/2020\|EPI_ISL_441617\|202 | ···················· | ···················· | ···················· | ···················· | ···················· | ···················· |
| B | 5287 | hCoV-19/England/CAMB-7C5F1/2020\|EPI_ISL_441619\|202 | ···················· | ···················· | ···················· | ···················· | ···················· | ···················· |
| B | 5288 | hCoV-19/England/CAMB-7F558/2020\|EPI_ISL_442910\|202 | ···················· | ···················· | ···················· | ···················· | ···················· | ···················· |
| B | 5289 | hCoV-19/England/CAMB-809D0/2020\|EPI_ISL_442912\|202 | ···················· | ···················· | ···················· | ···················· | ···················· | ···················· |
| B | 5290 | hCoV-19/England/CAMB-80C7D/2020\|EPI_ISL_442913\|202 | ···················· | ···················· | ···················· | ···················· | ···················· | ···················· |
| B | 5291 | hCoV-19/England/CAMB-80BF8/2020\|EPI_ISL_442914\|202 | ···················· | ···················· | ···················· | ···················· | ···················· | ···················· |
| B | 5292 | hCoV-19/England/CAMB-80C40/2020\|EPI_ISL_442915\|202 | ···················· | ···················· | ···················· | ···················· | ···················· | ···················· |
| B | 5293 | hCoV-19/England/CAMB-7F01B/2020\|EPI_ISL_442916\|202 | ···················· | ···················· | ···················· | ···················· | ···················· | ···················· |
| B | 5294 | hCoV-19/England/CAMB-7EB39/2020\|EPI_ISL_442917\|202 | ···················· | ···················· | ···················· | ···················· | ···················· | ···················· |
| B | 5295 | hCoV-19/England/CAMB-7F242/2020\|EPI_ISL_442918\|202 | ···················· | ···················· | ···················· | ···················· | ···················· | ···················· |
| B | 5296 | hCoV-19/England/CAMB-7F36D/2020\|EPI_ISL_442919\|202 | ···················· | ···················· | ···················· | ···················· | ···················· | ···················· |
| B | 5297 | hCoV-19/England/CAMB-7EDCA/2020\|EPI_ISL_442920\|202 | ···················· | ···················· | ···················· | ···················· | ···················· | ···················· |
| B | 5298 | hCoV-19/England/CAMB-8092B/2020\|EPI_ISL_442922\|202 | ···················· | ···················· | ···················· | ···················· | ···················· | ···················· |
| B | 5299 | hCoV-19/England/CAMB-7EF79/2020\|EPI_ISL_442923\|202 | ···················· | ···················· | ···················· | ···················· | ···················· | ···················· |
| B | 5300 | hCoV-19/England/CAMB-7F29D/2020\|EPI_ISL_442926\|202 | ···················· | ···················· | ···················· | ···················· | ···················· | ···················· |
| B | 5301 | hCoV-19/England/CAMB-81632/2020\|EPI_ISL_442927\|202 | ···················· | ···················· | ···················· | ···················· | ···················· | ···················· |
| B | 5302 | hCoV-19/England/CAMB-80C04/2020\|EPI_ISL_442940\|202 | ···················· | ···················· | ···················· | ···················· | ···················· | ···················· |
| B | 5303 | hCoV-19/England/CAMB-7F2F7/2020\|EPI_ISL_442943\|202 | ···················· | ···················· | ···················· | ···················· | ···················· | ···················· |
| B | 5304 | hCoV-19/England/CAMB-7F3E5/2020\|EPI_ISL_442944\|202 | ···················· | ···················· | ···················· | ···················· | ···················· | ···················· |
| B | 5305 | hCoV-19/England/CAMB-7EF2E/2020\|EPI_ISL_442945\|202 | ···················· | ···················· | ···················· | ···················· | ···················· | ···················· |
| B | 5306 | hCoV-19/England/CAMB-7F45B/2020\|EPI_ISL_442947\|202 | ···················· | ···················· | ···················· | ···················· | ···················· | ···················· |
| B | 5307 | hCoV-19/USA/CA-SR0104/2020\|EPI_ISL_445101\|2020-03- | ···················· | ···················· | ···················· | ···················· | ···················· | ···················· |
| B | 5308 | hCoV-19/USA/CA-SR0106/2020\|EPI_ISL_445102\|2020-03- | ···················· | ···················· | ···················· | ···················· | ···················· | ···················· |
| B | 5309 | hCoV-19/USA/CA-SR0111/2020\|EPI_ISL_445105\|2020-03- | ···················· | ···················· | ···················· | ···················· | ···················· | ···················· |
| B | 5310 | hCoV-19/USA/CA-SR0115/2020\|EPI_ISL_445107\|2020-03- | ···················· | ···················· | ···················· | ···················· | ···················· | ···················· |
| B | 5311 | hCoV-19/USA/CA-SR0117/2020\|EPI_ISL_445108\|2020-03- | ···················· | ···················· | ···················· | ···················· | ···················· | ···················· |
| B | 5312 | hCoV-19/USA/CA-SR0118/2020\|EPI_ISL_445109\|2020-03- | ···················· | ···················· | ···················· | ···················· | ···················· | ···················· |
| B | 5313 | hCoV-19/USA/CA-SR0119/2020\|EPI_ISL_445110\|2020-03- | ···················· | ···················· | ···················· | ···················· | ···················· | ···················· |
| B | 5314 | hCoV-19/USA/CA-SR0120/2020\|EPI_ISL_445111\|2020-03- | ···················· | ···················· | ···················· | ···················· | ···················· | ···················· |
| B | 5315 | hCoV-19/USA/CA-SR0128/2020\|EPI_ISL_445113\|2020-03- | ···················· | ···················· | ···················· | ···················· | ···················· | ···················· |
| B | 5316 | hCoV-19/USA/CA-SR0130/2020\|EPI_ISL_445114\|2020-03- | ···················· | ···················· | ···················· | ···················· | ···················· | ···················· |
| B | 5317 | hCoV-19/USA/CA-SR0132/2020\|EPI_ISL_445115\|2020-03- | ···················· | ···················· | ···················· | ···················· | ···················· | ···················· |
| B | 5318 | hCoV-19/USA/CA-SR0135/2020\|EPI_ISL_445116\|2020-03- | ···················· | ···················· | ···················· | ···················· | ···················· | ···················· |
| B | 5319 | hCoV-19/USA/CA-SR0136/2020\|EPI_ISL_445117\|2020-03- | ···················· | ···················· | ···················· | ···················· | ···················· | ···················· |
| B | 5320 | hCoV-19/USA/CA-SR0217/2020\|EPI_ISL_445118\|2020-05- | ···················· | ·A·················· | ···················· | ···················· | ···················· | ···················· |
| B | 5321 | hCoV-19/USA/LA-SR0264/2020\|EPI_ISL_445119\|2020-03- | ···················· | ···················· | ···················· | ···················· | ···················· | ···················· |
| B | 5322 | hCoV-19/USA/LA-SR0265/2020\|EPI_ISL_445120\|2020-04- | ···················· | ···················· | ···················· | ···················· | ···················· | ···················· |
| B | 5323 | hCoV-19/USA/LA-SR0266/2020\|EPI_ISL_445121\|2020-03- | ···················· | ···················· | ···················· | ···················· | ···················· | ···················· |
| B | 5324 | hCoV-19/USA/LA-SR0267/2020\|EPI_ISL_445122\|2020-03- | ···················· | ···················· | ···················· | ···················· | ···················· | ···················· |
| B | 5325 | hCoV-19/USA/LA-SR0268/2020\|EPI_ISL_445123\|2020-04- | ···················· | ···················· | ···················· | ···················· | ···················· | ···················· |
| B | 5326 | hCoV-19/USA/LA-SR0269/2020\|EPI_ISL_445124\|2020-04- | ···················· | ···················· | ···················· | ···················· | ···················· | ···················· |
| B | 5327 | hCoV-19/USA/LA-SR0270/2020\|EPI_ISL_445125\|2020-04- | ···················· | ···················· | ···················· | ···················· | ···················· | ···················· |
| B | 5328 | hCoV-19/USA/LA-SR0271/2020\|EPI_ISL_445126\|2020-03- | ···················· | ···················· | ···················· | ···················· | ···················· | ···················· |
| B | 5329 | hCoV-19/USA/LA-SR0272/2020\|EPI_ISL_445127\|2020-04- | ···················· | ···················· | ···················· | ···················· | ···················· | ···················· |
| B | 5330 | hCoV-19/USA/LA-SR0273/2020\|EPI_ISL_445128\|2020-04- | ···················· | ···················· | ···················· | ···················· | ···················· | ···················· |
| B | 5331 | hCoV-19/USA/LA-SR0274/2020\|EPI_ISL_445129\|2020-04- | ···················· | ···················· | ···················· | ···················· | ···················· | ···················· |
| B | 5332 | hCoV-19/USA/LA-SR0275/2020\|EPI_ISL_445130\|2020-03- | ···················· | ···················· | ···················· | ···················· | ···················· | ···················· |
| B | 5333 | hCoV-19/USA/LA-SR0277/2020\|EPI_ISL_445132\|2020-04- | ···················· | ···················· | ···················· | ···················· | ···················· | ···················· |
| B | 5334 | hCoV-19/USA/LA-SR0278/2020\|EPI_ISL_445133\|2020-04- | ···················· | ···················· | ···················· | ···················· | ···················· | ···················· |
| B | 5335 | hCoV-19/USA/LA-SR0279/2020\|EPI_ISL_445134\|2020-03- | ···················· | ···················· | ···················· | ···················· | ···················· | ···················· |
| B | 5336 | hCoV-19/USA/LA-SR0280/2020\|EPI_ISL_445135\|2020-04- | ···················· | ···················· | ···················· | ···················· | ···················· | ···················· |
| B | 5337 | hCoV-19/USA/LA-SR0281/2020\|EPI_ISL_445136\|2020-03- | ···················· | ···················· | ···················· | ···················· | ···················· | ···················· |
| B | 5338 | hCoV-19/USA/LA-SR0282/2020\|EPI_ISL_445137\|2020-04- | ···················· | ···················· | ···················· | ···················· | ···················· | ···················· |
| B | 5339 | hCoV-19/USA/LA-SR0283/2020\|EPI_ISL_445138\|2020-03- | ···················· | ···················· | ···················· | ···················· | ···················· | ···················· |
| B | 5340 | hCoV-19/USA/LA-SR0284/2020\|EPI_ISL_445139\|2020-04- | ···················· | ···················· | ···················· | ···················· | ···················· | ···················· |
| B | 5341 | hCoV-19/USA/LA-SR0285/2020\|EPI_ISL_445140\|2020-04- | ···················· | ···················· | ···················· | ···················· | ···················· | ···················· |
| B | 5342 | hCoV-19/USA/LA-SR0286/2020\|EPI_ISL_445141\|2020-03- | ···················· | ···················· | ···················· | ···················· | ···················· | ···················· |
| B | 5343 | hCoV-19/USA/LA-SR0288/2020\|EPI_ISL_445143\|2020-04- | ···················· | ···················· | ···················· | ···················· | ···················· | ···················· |
| B | 5344 | hCoV-19/USA/LA-SR0289/2020\|EPI_ISL_445144\|2020-03- | ···················· | ···················· | ···················· | ···················· | ···················· | ···················· |
| B | 5345 | hCoV-19/USA/LA-SR0290/2020\|EPI_ISL_445145\|2020-04- | ···················· | ···················· | ···················· | ···················· | ···················· | ···················· |

| | | | | | | | | |
|---|---|---|---|---|---|---|---|---|
| B | 5346 hCoV-19/USA/LA-SR0291/2020\|EPI_ISL_445146\|2020-04- | ···················· | ···················· | ···················· | ···················· | ···················· | ···················· | |
| B | 5347 hCoV-19/USA/LA-SR0292/2020\|EPI_ISL_445147\|2020-04- | ···················· | ···················· | ···················· | ···················· | ···················· | ···················· | |
| B | 5348 hCoV-19/USA/LA-SR0293/2020\|EPI_ISL_445148\|2020-04- | ···················· | ···················· | ···················· | ···················· | ···················· | ···················· | |
| B | 5349 hCoV-19/USA/LA-SR0294/2020\|EPI_ISL_445149\|2020-04- | ···················· | ···················· | ···················· | ···················· | ···················· | ···················· | |
| B | 5350 hCoV-19/Wales/PHWC-3122E/2020\|EPI_ISL_446423\|2020- | ···················· | ···················· | ···················· | ···················· | ···················· | ···················· | |
| B | 5351 hCoV-19/Wales/PHWC-3124C/2020\|EPI_ISL_446425\|2020- | ···················· | ···················· | ···················· | ···················· | ···················· | ···················· | |
| B | 5352 hCoV-19/Wales/PHWC-31358/2020\|EPI_ISL_446435\|2020- | ···················· | ···················· | ···················· | ···················· | ···················· | ···················· | |
| B | 5353 hCoV-19/Wales/PHWC-313EF/2020\|EPI_ISL_446441\|2020- | ···················· | ····A··············· | ···················· | ···················· | ···················· | ···················· | |
| B | 5354 hCoV-19/Wales/PHWC-313FE/2020\|EPI_ISL_446442\|2020- | ···················· | ···················· | ···················· | ···················· | ···················· | ···················· | |
| B | 5355 hCoV-19/Wales/PHWC-314FB/2020\|EPI_ISL_446455\|2020- | ···················· | ···················· | ···················· | ···················· | ···················· | ···················· | |
| B | 5356 hCoV-19/Wales/PHWC-31561/2020\|EPI_ISL_446461\|2020- | ···················· | ···················· | ···················· | ···················· | ···················· | ···················· | |
| B | 5357 hCoV-19/Wales/PHWC-3159E/2020\|EPI_ISL_446464\|2020- | ···················· | ···················· | ···················· | ···················· | ···················· | ···················· | |
| B | 5358 hCoV-19/Wales/PHWC-315DA/2020\|EPI_ISL_446466\|2020- | ···················· | ···················· | ···················· | ···················· | ···················· | ···················· | |
| B | 5359 hCoV-19/Wales/PHWC-316B9/2020\|EPI_ISL_446473\|2020- | ···················· | ···················· | ···················· | ···················· | ···················· | ···················· | |
| B | 5360 hCoV-19/Wales/PHWC-316D7/2020\|EPI_ISL_446474\|2020- | ···················· | ···················· | ···················· | ···················· | ···················· | ···················· | |
| B | 5361 hCoV-19/Wales/PHWC-316E6/2020\|EPI_ISL_446475\|2020- | ···················· | ···················· | ···················· | ···················· | ···················· | ···················· | |
| B | 5362 hCoV-19/Wales/PHWC-316F5/2020\|EPI_ISL_446476\|2020- | ···················· | ···················· | ···················· | ···················· | ···················· | ···················· | |
| B | 5363 hCoV-19/Wales/PHWC-31701/2020\|EPI_ISL_446477\|2020- | ···················· | ···················· | ···················· | ···················· | ···················· | ···················· | |
| B | 5364 hCoV-19/Wales/PHWC-3224B/2020\|EPI_ISL_446621\|2020- | ···················· | ···················· | ···················· | ···················· | ···················· | ···················· | |
| B | 5365 hCoV-19/Wales/PHWC-32269/2020\|EPI_ISL_446623\|2020- | ···················· | ···················· | ···················· | ···················· | ···················· | ···················· | |
| B | 5366 hCoV-19/France/IDF-40003KA/2020\|EPI_ISL_447724\|202 | ···················· | ···················· | ···················· | ···················· | ···················· | ···················· | |
| B | 5367 hCoV-19/France/IDF-10007LJ/2020\|EPI_ISL_447725\|202 | ···················· | ···················· | ···················· | ···················· | ···················· | ···················· | |
| B | 5368 hCoV-19/France/IDF-10028CD/2020\|EPI_ISL_447726\|202 | ···················· | ···················· | ···················· | ···················· | ···················· | ···················· | |
| B | 5369 hCoV-19/France/IDF-10006HC/2020\|EPI_ISL_447727\|202 | ···················· | ···················· | ···················· | ···················· | ···················· | ···················· | |
| B | 5370 hCoV-19/France/IDF-10053BP/2020\|EPI_ISL_447728\|202 | ···················· | ···················· | ···················· | ···················· | ···················· | ···················· | |
| B | 5371 hCoV-19/France/IDF-10064DR/2020\|EPI_ISL_447729\|202 | ···················· | ···················· | ···················· | ···················· | ···················· | ···················· | |
| B | 5372 hCoV-19/France/IDF-10065EE/2020\|EPI_ISL_447730\|202 | ···················· | ···················· | ···················· | ···················· | ···················· | ···················· | |
| B | 5373 hCoV-19/France/IDF-10070SK/2020\|EPI_ISL_447731\|202 | ···················· | ···················· | ···················· | ···················· | ···················· | ···················· | |
| B | 5374 hCoV-19/France/IDF-50006PT/2020\|EPI_ISL_447732\|202 | ···················· | ···················· | ···················· | ···················· | ···················· | ···················· | |
| B | 5375 hCoV-19/France/IDF-40002VJ/2020\|EPI_ISL_447733\|202 | ···················· | ···················· | ···················· | ···················· | ···················· | ···················· | |
| B | 5376 hCoV-19/India/un-OUMRK1090/2020\|EPI_ISL_447902\|202 | ···················· | ···················· | ···················· | ···················· | ···················· | ···················· | |
| B | 5377 hCoV-19/Scotland/CVR473/2020\|EPI_ISL_433302\|2020-0 | ···················· | ···················A | ···················· | ···················· | ···················· | ···················· | 1 |
| B | 5378 hCoV-19/Scotland/CVR487/2020\|EPI_ISL_433303\|2020-0 | ···················· | ·A·················· | ···················· | ···················· | ···················· | ···················· | |
| B | 5379 hCoV-19/Scotland/CVR491/2020\|EPI_ISL_433304\|2020-0 | ···················· | ···················· | ···················· | ···················· | ···················· | ···················· | |
| B | 5380 hCoV-19/Scotland/CVR495/2020\|EPI_ISL_433305\|2020-0 | ···················· | ·A·················· | ···················· | ···················· | ···················· | ···················· | |
| B | 5381 hCoV-19/Scotland/CVR507/2020\|EPI_ISL_433307\|2020-0 | ···················· | ·A·················· | ···················· | ···················· | ···················· | ···················· | |
| B | 5382 hCoV-19/Scotland/CVR583/2020\|EPI_ISL_433313\|2020-0 | ···················· | ···················· | ···················· | ···················· | ···················· | ···················· | |
| B | 5383 hCoV-19/Scotland/CVR599/2020\|EPI_ISL_433314\|2020-0 | ···················· | ·A·················· | ···················· | ···················· | ···················· | ···················· | |
| B | 5384 hCoV-19/Scotland/CVR607/2020\|EPI_ISL_433316\|2020-0 | ···················· | ·A·················· | ···················· | ···················· | ···················· | ···················· | |
| B | 5385 hCoV-19/Scotland/CVR663/2020\|EPI_ISL_433321\|2020-0 | ···················· | ···················· | ···················· | ···················· | ···················· | ···················· | |
| B | 5386 hCoV-19/Scotland/CVR672/2020\|EPI_ISL_433323\|2020-0 | ···················· | ···················· | ···················· | ···················· | ···················· | ···················· | |
| B | 5387 hCoV-19/Scotland/CVR681/2020\|EPI_ISL_433324\|2020-0 | ···················· | ···················· | ···················· | ···················· | ···················· | ···················· | |
| B | 5388 hCoV-19/USA/WI-UW-266/2020\|EPI_ISL_434612\|2020-04- | ···················· | ···················· | ···················· | ···················· | ···················· | ···················· | |
| B | 5389 hCoV-19/USA/WI-UW-267/2020\|EPI_ISL_434613\|2020-04- | ···················· | ···················· | ···················· | ···················· | ···················· | ···················· | |
| B | 5390 hCoV-19/USA/WI-UW-268/2020\|EPI_ISL_434614\|2020-04- | ···················· | ···················· | ···················· | ···················· | ···················· | ···················· | |
| B | 5391 hCoV-19/USA/WI-UW-269/2020\|EPI_ISL_434615\|2020-04- | ···················· | ···················· | ···················· | ···················· | ···················· | ···················· | |
| B | 5392 hCoV-19/France/OCC-1/2020\|EPI_ISL_434616\|2020-03 | ···T················ | ···················· | ···················· | ···················· | ···················· | ···················· | |
| B | 5393 hCoV-19/France/OCC-2/2020\|EPI_ISL_434617\|2020-03 | ···················· | ···················· | ···················· | ···················· | ···················· | ···················· | |
| B | 5394 hCoV-19/France/OCC-3/2020\|EPI_ISL_434618\|2020-03 | ···T················ | ···················· | ···················· | ···················· | ···················· | ···················· | |
| B | 5395 hCoV-19/France/OCC-4/2020\|EPI_ISL_434619\|2020-03 | ···················· | ···················· | ···················· | ···················· | ···················· | ···················· | |
| B | 5396 hCoV-19/France/OCC-5/2020\|EPI_ISL_434620\|2020-03 | ···················· | ···················· | ···················· | ···················· | ···················· | ···················· | |
| B | 5397 hCoV-19/France/OCC-6/2020\|EPI_ISL_434621\|2020-03 | ···················· | ···················· | ···················· | ···················· | ···················· | ···················· | |
| B | 5398 hCoV-19/France/OCC-7/2020\|EPI_ISL_434622\|2020-03 | ···················· | ···················· | ···················· | ···················· | ···················· | ···················· | |
| B | 5399 hCoV-19/France/OCC-8/2020\|EPI_ISL_434623\|2020-03 | ···················· | ···················· | ···················· | ···················· | ···················· | ···················· | |
| B | 5400 hCoV-19/France/OCC-9/2020\|EPI_ISL_434624\|2020-03 | ···················· | ···················· | ···················· | ···················· | ···················· | ···················· | |
| B | 5401 hCoV-19/France/OCC-10/2020\|EPI_ISL_434625\|2020-03 | ···················· | ···················· | ···················· | ···················· | ···················· | ···················· | |
| B | 5402 hCoV-19/France/OCC-11/2020\|EPI_ISL_434626\|2020-03- | ···················· | ···················· | ···················· | ···················· | ···················· | ···················· | |
| B | 5403 hCoV-19/France/OCC-12/2020\|EPI_ISL_434627\|2020-03- | ···················· | ···················· | ···················· | ···················· | ···················· | ···················· | |
| B | 5404 hCoV-19/France/OCC-13/2020\|EPI_ISL_434628\|2020-04- | ···················· | ···················· | ···················· | ···················· | ···················· | ···················· | |
| B | 5405 hCoV-19/France/OCC-14/2020\|EPI_ISL_434629\|2020-04- | ···················· | ···················· | ···················· | ···················· | ···················· | ···················· | |
| B | 5406 hCoV-19/France/OCC-15/2020\|EPI_ISL_434630\|2020-04- | ···················· | ···················· | ···················· | ···················· | ···················· | ···················· | |
| B | 5407 hCoV-19/France/OCC-16/2020\|EPI_ISL_434631\|2020-04- | ···················· | ···················· | ···················· | ···················· | ···················· | ···················· | |
| B | 5408 hCoV-19/France/OCC-17/2020\|EPI_ISL_434632\|2020-04- | ···················· | ···················· | ···················· | ···················· | ···················· | ···················· | |
| B | 5409 hCoV-19/France/OCC-18/2020\|EPI_ISL_434633\|2020-04- | ···················· | ···················· | ···················· | ···················· | ···················· | ···················· | |
| B | 5410 hCoV-19/France/OCC-19/2020\|EPI_ISL_434634\|2020-04- | ···················· | ···················· | ···················· | ···················· | ···················· | ···················· | |
| B | 5411 hCoV-19/France/OCC-20/2020\|EPI_ISL_434635\|2020-04- | ···················· | ···················· | ···················· | ···················· | ···················· | ···················· | |
| B | 5412 hCoV-19/USA/FL-UF1/2020\|EPI_ISL_434636\|2020-03-10 | ···················· | ···················· | ···················· | ···················· | ···················· | ···················· | |
| B | 5413 hCoV-19/USA/FL-UF2/2020\|EPI_ISL_434637\|2020-03-27 | ···················· | ···················· | ···················· | ···················· | ···················· | ···················· | |
| B | 5414 hCoV-19/USA/MD-HP00041/2020\|EPI_ISL_434638\|2020-03 | ···················· | ···················· | ···················· | ···················· | ···················· | ···················· | |
| B | 5415 hCoV-19/USA/MD-HP00044/2020\|EPI_ISL_434640\|2020-03 | ···················· | ···················· | ···················· | ···················· | ···················· | ···················· | |
| B | 5416 hCoV-19/Sweden/20-08682/2020\|EPI_ISL_434641\|2020-0 | ···················· | ···················· | ···················· | ···················· | ···················· | ···················· | |
| B | 5417 hCoV-19/Sweden/20-08686/2020\|EPI_ISL_434642\|2020-0 | ···················· | ···················· | ···················· | ···················· | ···················· | ···················· | |
| B | 5418 hCoV-19/Sweden/20-05448/2020\|EPI_ISL_434643\|2020-0 | ···················· | ···················· | ···················· | ···················· | ···················· | ···················· | |
| B | 5419 hCoV-19/Sweden/20-05469/2020\|EPI_ISL_434644\|2020-0 | ···················· | ···················· | ···················· | ···················· | ···················· | ···················· | |
| B | 5420 hCoV-19/Sweden/20-05627/2020\|EPI_ISL_434645\|2020-0 | ···················· | ···················· | ···················· | ···················· | ···················· | ···················· | |
| B | 5421 hCoV-19/Sweden/20-05653/2020\|EPI_ISL_434646\|2020-0 | ···················· | ···················· | ···················· | ···················· | ···················· | ···················· | |
| B | 5422 hCoV-19/Sweden/20-05721/2020\|EPI_ISL_434647\|2020-0 | ···················· | ···················· | ···················· | ···················· | ···················· | ···················· | |
| B | 5423 hCoV-19/Sweden/20-05847/2020\|EPI_ISL_434648\|2020-0 | ···················· | ···················· | ···················· | ···················· | ···················· | ···················· | |

| | | | | | | | |
|---|---|---|---|---|---|---|---|
| B | 5424 hCoV-19/Sweden/20-05856/2020\|EPI_ISL_434649\|2020-0 | ···················· | ···················· | ····················· | ···················· | ···················· | ···················· |
| B | 5425 hCoV-19/Sweden/20-06071/2020\|EPI_ISL_434650\|2020-0 | ···················· | ···················· | ····················· | ···················· | ···················· | ···················· |
| B | 5426 hCoV-19/Sweden/20-06306/2020\|EPI_ISL_434651\|2020-0 | ···················· | ···················· | ····················· | ···················· | ···················· | ···················· |
| B | 5427 hCoV-19/Sweden/20-06308/2020\|EPI_ISL_434652\|2020-0 | ···················· | ···················· | ····················· | ···················· | ···················· | ···················· |
| B | 5428 hCoV-19/Sweden/20-06376/2020\|EPI_ISL_434653\|2020-0 | ···················· | ···················· | ····················· | ···················· | ···················· | ···················· |
| B | 5429 hCoV-19/Sweden/20-06383/2020\|EPI_ISL_434654\|2020-0 | ···················· | ···················· | ····················· | ···················· | ···················· | ···················· |
| B | 5430 hCoV-19/Sweden/20-06468/2020\|EPI_ISL_434655\|2020-0 | ···················· | ···················· | ····················· | ···················· | ···················· | ···················· |
| B | 5431 hCoV-19/Austria/CeMM0273/2020\|EPI_ISL_438100\|2020- | ···················· | ···················· | ····················· | ···················· | ···················· | ···················· |
| B | 5432 hCoV-19/Austria/CeMM0274/2020\|EPI_ISL_438101\|2020- | ···················· | ···················· | ····················· | ···················· | ···················· | ···················· |
| B | 5433 hCoV-19/Austria/CeMM0275/2020\|EPI_ISL_438102\|2020- | ···················· | ···················· | ····················· | ···················· | ···················· | ···················· |
| B | 5434 hCoV-19/Austria/CeMM0276/2020\|EPI_ISL_438103\|2020- | ···················· | ···················· | ····················· | ···················· | ···················· | ···················· |
| B | 5435 hCoV-19/Austria/CeMM0277/2020\|EPI_ISL_438104\|2020- | ···················· | ···················· | ····················· | ···················· | ···················· | ···················· |
| B | 5436 hCoV-19/Austria/CeMM0278/2020\|EPI_ISL_438105\|2020- | ···················· | ···················· | ····················· | ···················· | ···················· | ···················· |
| B | 5437 hCoV-19/Austria/CeMM0279/2020\|EPI_ISL_438106\|2020- | ···················· | ···················· | ····················· | ···················· | ···················· | ···················· |
| B | 5438 hCoV-19/Austria/CeMM0280/2020\|EPI_ISL_438107\|2020- | ···················· | ···················· | ····················· | ···················· | ···················· | ···················· |
| B | 5439 hCoV-19/Austria/CeMM0281/2020\|EPI_ISL_438108\|2020- | ···················· | ···················· | ····················· | ···················· | ···················· | ···················· |
| B | 5440 hCoV-19/Austria/CeMM0283/2020\|EPI_ISL_438109\|2020- | ···················· | ···················· | ····················· | ···················· | ···················· | ···················· |
| B | 5441 hCoV-19/Austria/CeMM0284/2020\|EPI_ISL_438110\|2020- | ···················· | ···················· | ····················· | ···················· | ···················· | ···················· |
| B | 5442 hCoV-19/Austria/CeMM0285/2020\|EPI_ISL_438111\|2020- | ···················· | ···················· | ····················· | ···················· | ···················· | ···················· |
| B | 5443 hCoV-19/Austria/CeMM0287/2020\|EPI_ISL_438112\|2020- | ···················· | ···················· | ····················· | ···················· | ···················· | ···················· |
| B | 5444 hCoV-19/Austria/CeMM0288/2020\|EPI_ISL_438113\|2020- | ···················· | ···················· | ····················· | ···················· | ···················· | ···················· |
| B | 5445 hCoV-19/Austria/CeMM0290/2020\|EPI_ISL_438114\|2020- | ···················· | ···················· | ····················· | ···················· | ···················· | ···················· |
| B | 5446 hCoV-19/Austria/CeMM0291/2020\|EPI_ISL_438115\|2020- | ···················· | ···················· | ····················· | ···················· | ···················· | ···················· |
| B | 5447 hCoV-19/Austria/CeMM0292/2020\|EPI_ISL_438116\|2020- | ···················· | ···················· | ····················· | ···················· | ···················· | ···················· |
| B | 5448 hCoV-19/Austria/CeMM0294/2020\|EPI_ISL_438117\|2020- | ···················· | ···················· | ····················· | ···················· | ···················· | ···················· |
| B | 5449 hCoV-19/Austria/CeMM0295/2020\|EPI_ISL_438118\|2020- | ···················· | ···················· | ····················· | ···················· | ···················· | ·N·················· |
| B | 5450 hCoV-19/Austria/CeMM0296/2020\|EPI_ISL_438119\|2020- | ···················· | ···················· | ····················· | ···················· | ···················· | ···················· |
| B | 5451 hCoV-19/Austria/CeMM0357/2020\|EPI_ISL_438120\|2020- | ···················· | ···················· | ····················· | ···················· | ···················· | ···················· |
| B | 5452 hCoV-19/Austria/CeMM0358/2020\|EPI_ISL_438121\|2020- | ···················· | ···················· | ····················· | ···················· | ···················· | ···················· |
| B | 5453 hCoV-19/Austria/CeMM0359/2020\|EPI_ISL_438122\|2020- | ···················· | ···················· | ····················· | ···················· | ···················· | ···················· |
| B | 5454 hCoV-19/Austria/CeMM0360/2020\|EPI_ISL_438123\|2020- | ···················· | ···················· | ··········M·········· | ···················· | ···················· | ···················· |
| B | 5455 hCoV-19/Austria/CeMM0361/2020\|EPI_ISL_438124\|2020- | ···················· | ···················· | ····················· | ···················· | ···················· | ···················· |
| B | 5456 hCoV-19/Austria/CeMM0362/2020\|EPI_ISL_438125\|2020- | ···················· | ···················· | ····················· | ···················· | ···················· | ···················· |
| B | 5457 hCoV-19/Austria/CeMM0363/2020\|EPI_ISL_438126\|2020- | ···················· | ···················· | ····················· | ···················· | ···················· | ···················· |
| B | 5458 hCoV-19/Austria/CeMM0364/2020\|EPI_ISL_438127\|2020- | ···················· | ···················· | ····················· | ···················· | ···················· | ···················· |
| B | 5459 hCoV-19/Austria/CeMM0365/2020\|EPI_ISL_438128\|2020- | ···················· | ···················· | ··Y·················· | ···················· | ···················· | ···················· |
| B | 5460 hCoV-19/India/TG-GMC-RR1191/2020\|EPI_ISL_438138\|20 | ···················· | ···················· | ····················· | ···················· | ···················· | ···················· |
| B | 5461 hCoV-19/India/TG-GMC-RK1090/2020\|EPI_ISL_438139\|20 | ···················· | ···················· | ····················· | ···················· | ···················· | ···················· |
| B | 5462 hCoV-19/USA/WA-S587/2020\|EPI_ISL_438140\|2020-04-01 | ···················· | ···················· | ····················· | ···················· | ···················· | ···················· |
| B | 5463 hCoV-19/USA/WA-S588/2020\|EPI_ISL_438141\|2020-04-02 | ···················· | ···················· | ····················· | ···················· | ···················· | ···················· |
| B | 5464 hCoV-19/USA/WA-S589/2020\|EPI_ISL_438142\|2020-04-04 | ···················· | ···················· | ····················· | ···················· | ···················· | ···················· |
| B | 5465 hCoV-19/USA/WA-S590/2020\|EPI_ISL_438143\|2020-04-04 | ···················· | ···················· | ····················· | ···················· | ···················· | ···················· |
| B | 5466 hCoV-19/USA/WA-S591/2020\|EPI_ISL_438144\|2020-04-04 | ···················· | ···················· | ····················· | ···················· | ···················· | ···················· |
| B | 5467 hCoV-19/USA/WA-S592/2020\|EPI_ISL_438145\|2020-04-04 | ···················· | ···················· | ····················· | ···················· | ···················· | ···················· |
| B | 5468 hCoV-19/USA/WA-S594/2020\|EPI_ISL_438147\|2020-04-12 | ···················· | ···················· | ····················· | ···················· | ···················· | ···················· |
| B | 5469 hCoV-19/USA/WA-S595/2020\|EPI_ISL_438148\|2020-04-12 | ···················· | ···················· | ····················· | ···················· | ···················· | ···················· |
| B | 5470 hCoV-19/USA/WA-S596/2020\|EPI_ISL_438149\|2020-04-13 | ···················· | ···················· | ····················· | ···················· | ···················· | ···················· |
| B | 5471 hCoV-19/USA/WA-S607/2020\|EPI_ISL_438160\|2020-04-20 | ···················· | ···················· | ····················· | ···················· | ···················· | ···················· |
| B | 5472 hCoV-19/USA/WA-S608/2020\|EPI_ISL_438161\|2020-04-19 | ···················· | ···················· | ····················· | ···················· | ···················· | ···················· |
| B | 5473 hCoV-19/USA/WA-S609/2020\|EPI_ISL_438162\|2020-04-23 | ···················· | ···················· | ····················· | ···················· | ···················· | ···················· |
| B | 5474 hCoV-19/USA/WA-S610/2020\|EPI_ISL_438163\|2020-04-23 | ···················· | ···················· | ····················· | ···················· | ···················· | ···················· |
| B | 5476 hCoV-19/USA/WA-S612/2020\|EPI_ISL_438165\|2020-04-23 | ···················· | ···················· | ····················· | ···················· | ···················· | ···················· |
| B | 5477 hCoV-19/USA/WA-S613/2020\|EPI_ISL_438166\|2020-04-23 | ···················· | ···················· | ····················· | ···················· | ···················· | ···················· |
| B | 5478 hCoV-19/USA/WA-S614/2020\|EPI_ISL_438167\|2020-04-23 | ···················· | ···················· | ····················· | ···················· | ···················· | ···················· |
| B | 5479 hCoV-19/USA/WA-S615/2020\|EPI_ISL_438168\|2020-04-23 | ···················· | ···················· | ····················· | ···················· | ···················· | ···················· |
| B | 5480 hCoV-19/USA/WA-S616/2020\|EPI_ISL_438169\|2020-04-23 | ···················· | ···················· | ····················· | ···················· | ···················· | ···················· |
| B | 5481 hCoV-19/USA/WA-S617/2020\|EPI_ISL_438170\|2020-04-25 | ···················· | ···················· | ····················· | ···················· | ···················· | ···················· |
| B | 5482 hCoV-19/USA/WA-S618/2020\|EPI_ISL_438171\|2020-04-26 | ···················· | ···················· | ····················· | ···················· | ···················· | ···················· |
| B | 5483 hCoV-19/USA/WA-S619/2020\|EPI_ISL_438172\|2020-04-26 | ···················· | ···················· | ····················· | ···················· | ···················· | ···················· |
| B | 5484 hCoV-19/USA/WA-S620/2020\|EPI_ISL_438173\|2020-04-26 | ···················· | ···················· | ····················· | ···················· | ···················· | ···················· |
| B | 5485 hCoV-19/USA/WA-S621/2020\|EPI_ISL_438174\|2020-04-27 | ···················· | ···················· | ····················· | ···················· | ···················· | ···················· |
| B | 5486 hCoV-19/USA/WA-S622/2020\|EPI_ISL_438175\|2020-04-29 | ···················· | ···················· | ····················· | ···················· | ···················· | ···················· |
| B | 5487 hCoV-19/USA/WA-S623/2020\|EPI_ISL_438176\|2020-04-18 | ···················· | ···················· | ····················· | ···················· | ···················· | ···················· |
| B | 5488 hCoV-19/USA/WA-S624/2020\|EPI_ISL_438177\|2020-04-18 | ···················· | ···················· | ····················· | ···················· | ···················· | ···················· |
| B | 5489 hCoV-19/USA/WA-S626/2020\|EPI_ISL_438179\|2020-04-18 | ···················· | ···················· | ····················· | ···················· | ···················· | ···················· |
| B | 5490 hCoV-19/England/CAMB-7767B/2020\|EPI_ISL_439430\|202 | ···················· | ···················· | ····················· | ···················· | ···················· | ···················· |
| B | 5491 hCoV-19/England/CAMB-77909/2020\|EPI_ISL_439431\|202 | ···················· | ···················· | ····················· | ···················· | ···················· | ···················· |
| B | 5492 hCoV-19/England/CAMB-760AB/2020\|EPI_ISL_439432\|202 | ···················· | ···················· | ····················· | ···················· | ···················· | ···················· |
| B | 5493 hCoV-19/England/CAMB-77B4F/2020\|EPI_ISL_439433\|202 | ···················· | ···················· | ····················· | ···················· | ···················· | ···················· |
| B | 5494 hCoV-19/England/CAMB-77110/2020\|EPI_ISL_439434\|202 | ···················· | ···················· | ····················· | ···················· | ···················· | ···················· |
| B | 5495 hCoV-19/England/CAMB-772FF/2020\|EPI_ISL_439436\|202 | ···················· | ···················· | ····················· | ···················· | ···················· | ···················· |
| B | 5496 hCoV-19/England/CAMB-7714D/2020\|EPI_ISL_439437\|202 | ···················· | ···················· | ····················· | ···················· | ···················· | ···················· |
| B | 5497 hCoV-19/England/CAMB-778C0/2020\|EPI_ISL_439438\|202 | ···················· | ···················· | ····················· | ···················· | ···················· | ···················· |
| B | 5498 hCoV-19/England/CAMB-77848/2020\|EPI_ISL_439439\|202 | ···················· | ···················· | ····················· | ···················· | ···················· | ···················· |
| B | 5499 hCoV-19/England/CAMB-77435/2020\|EPI_ISL_439440\|202 | ···················· | ···················· | ····················· | ···················· | ···················· | ···················· |
| B | 5500 hCoV-19/England/CAMB-778B1/2020\|EPI_ISL_439441\|202 | ···················· | ···················· | ····················· | ···················· | ···················· | ···················· |
| B | 5501 hCoV-19/England/CAMB-76102/2020\|EPI_ISL_439442\|202 | ···················· | ···················· | ····················· | ···················· | ···················· | ···················· |
| B | 5502 hCoV-19/England/CAMB-7768A/2020\|EPI_ISL_439443\|202 | ···················· | ···················· | ····················· | ···················· | ···················· | ···················· |

```
B 5503 hCoV-19/England/CAMB-77A7F/2020|EPI_ISL_439444|202 ···················· ···················· ····················· ···················· ···················· ····················
B 5504 hCoV-19/England/CAMB-76F44/2020|EPI_ISL_439446|202 ···················· ···················· ····················· ···················· ···················· ····················
B 5505 hCoV-19/England/CAMB-76F26/2020|EPI_ISL_439447|202 ···················· ···················· ····················· ···················· ···················· ····················
B 5506 hCoV-19/England/CAMB-76041/2020|EPI_ISL_439448|202 ···················· ···················· ····················· ···················· ···················· ····················
B 5507 hCoV-19/England/CAMB-776C6/2020|EPI_ISL_439449|202 ···················· ···················· ····················· ···················· ···················· ····················
B 5508 hCoV-19/England/CAMB-760E7/2020|EPI_ISL_439450|202 ···················· ············G······· ····················· ···················· ···················· ····················
B 5509 hCoV-19/England/CAMB-774AE/2020|EPI_ISL_439451|202 ···················· ···················· ····················· ···················· ···················· ····················
B 5510 hCoV-19/England/CAMB-76DA4/2020|EPI_ISL_439452|202 ···················· ···················· ····················· ···················· ···················· ····················
B 5511 hCoV-19/England/CAMB-77B8B/2020|EPI_ISL_439453|202 ···················· ···················· ····················· ···················· ···················· ····················
B 5512 hCoV-19/England/CAMB-77550/2020|EPI_ISL_439454|202 ···················· ···················· ····················· ···················· ···················· ····················
B 5513 hCoV-19/England/CAMB-7782A/2020|EPI_ISL_439455|202 ···················· ···················· ····················· ···················· ···················· ····················
B 5514 hCoV-19/England/CAMB-77392/2020|EPI_ISL_439456|202 ···················· ···················· ····················· ···················· ···················· ····················
B 5515 hCoV-19/England/CAMB-77778/2020|EPI_ISL_439457|202 ···················· ···················· ····················· ···················· ···················· ····················
B 5516 hCoV-19/England/CAMB-7717A/2020|EPI_ISL_439459|202 ···················· ···················· ····················· ···················· ···················· ····················
B 5517 hCoV-19/England/CAMB-77918/2020|EPI_ISL_439460|202 ···················· ···················· ····················· ···················· ···················· ····················
B 5518 hCoV-19/England/CAMB-7608D/2020|EPI_ISL_439461|202 ···················· ···················· ····················· ···················· ···················· ····················
B 5519 hCoV-19/England/CAMB-773CF/2020|EPI_ISL_439463|202 ···················· ···················· ····················· ···················· ···················· ····················
B 5520 hCoV-19/England/CAMB-77A9D/2020|EPI_ISL_439464|202 ···················· ···················· ····················· ···················· ···················· ····················
B 5521 hCoV-19/England/CAMB-7616C/2020|EPI_ISL_439465|202 ···················· ···················· ····················· ···················· ···················· ····················
B 5522 hCoV-19/England/CAMB-779AF/2020|EPI_ISL_439466|202 ···················· ···················· ····················· ···················· ···················· ····················
B 5523 hCoV-19/England/CAMB-7759C/2020|EPI_ISL_439468|202 ···················· ···················· ····················· ···················· ···················· ····················
B 5524 hCoV-19/England/CAMB-7614E/2020|EPI_ISL_439469|202 ···················· ···················· ····················· ···················· ···················· ····················
B 5525 hCoV-19/England/CAMB-77927/2020|EPI_ISL_439470|202 ···················· ···················· ····················· ···················· ···················· ····················
B 5526 hCoV-19/England/CAMB-765AC/2020|EPI_ISL_439471|202 ···················· ···················· ····················· ···················· ···················· ····················
B 5527 hCoV-19/England/CAMB-775D8/2020|EPI_ISL_439473|202 ···················· ···················· ····················· ···················· ···················· ····················
B 5528 hCoV-19/England/CAMB-7665E/2020|EPI_ISL_439475|202 ···················· ···················· ····················· ···················· ···················· ····················
B 5529 hCoV-19/England/CAMB-77963/2020|EPI_ISL_439476|202 ···················· ···················· ····················· ···················· ···················· ····················
B 5530 hCoV-19/England/CAMB-7763F/2020|EPI_ISL_439478|202 ···················· ···················· ····················· ···················· ···················· ····················
B 5531 hCoV-19/England/CAMB-77B6D/2020|EPI_ISL_439479|202 ···················· ···················· ····················· ···················· ···················· ····················
B 5532 hCoV-19/England/CAMB-774CC/2020|EPI_ISL_439490|202 ···················· ···················· ····················· ···················· ···················· ····················
B 5533 hCoV-19/England/CAMB-7666D/2020|EPI_ISL_439491|202 ···················· ···················· ····················· ···················· ···················· ····················
B 5534 hCoV-19/England/CAMB-771B6/2020|EPI_ISL_439492|202 ···················· ···················· ····················· ···················· ···················· ····················
B 5535 hCoV-19/England/CAMB-77480/2020|EPI_ISL_439494|202 ···················· ···················· ····················· ···················· ···················· ····················
B 5536 hCoV-19/England/CAMB-773ED/2020|EPI_ISL_439495|202 ···················· ···················· ····················· ···················· ···················· ····················
B 5537 hCoV-19/England/CAMB-7774B/2020|EPI_ISL_439496|202 ···················· ···················· ····················· ···················· ···················· ····················
B 5538 hCoV-19/England/CAMB-77462/2020|EPI_ISL_439498|202 ···················· ···················· ····················· ···················· ···················· ····················
B 5539 hCoV-19/England/CAMB-7730B/2020|EPI_ISL_439499|202 ···················· ···················· ····················· ···················· ···················· ····················
B 5540 hCoV-19/England/CAMB-7526A/2020|EPI_ISL_440411|202 ···················· ····A··············· ····················· ···················· ···················· ····················
B 5541 hCoV-19/England/CAMB-74F73/2020|EPI_ISL_440413|202 ···················· ···················· ····················· ···················· ···················· ····················
B 5542 hCoV-19/England/CAMB-75385/2020|EPI_ISL_440414|202 ···················· ···················· ····················· ···················· ···················· ····················
B 5543 hCoV-19/England/CAMB-752A6/2020|EPI_ISL_440417|202 ···················· ···················· ····················· ···················· ···················· ····················
B 5544 hCoV-19/England/CAMB-74F0A/2020|EPI_ISL_440418|202 ···················· ···················· ····················· ···················· ···················· ····················
B 5545 hCoV-19/England/CAMB-75394/2020|EPI_ISL_440419|202 ···················· ···················· ····················· ···················· ···················· ····················
B 5546 hCoV-19/England/CAMB-7517C/2020|EPI_ISL_440420|202 ···················· ···················· ····················· ···················· ···················· ····················
B 5547 hCoV-19/England/CAMB-7519A/2020|EPI_ISL_440421|202 ···················· ···················· ····················· ···················· ···················· ····················
B 5548 hCoV-19/England/CAMB-7522E/2020|EPI_ISL_440422|202 ···················· ···················· ····················· ···················· ···················· ····················
B 5549 hCoV-19/England/CAMB-756C8/2020|EPI_ISL_440426|202 ···················· ···················· ····················· ···················· ···················· ····················
B 5550 hCoV-19/England/CAMB-7532B/2020|EPI_ISL_440428|202 ···················· ···················· ····················· ···················· ···················· ····················
B 5551 hCoV-19/England/CAMB-753A3/2020|EPI_ISL_440429|202 ···················· ···················· ····················· ···················· ···················· ····················
B 5552 hCoV-19/England/CAMB-753EF/2020|EPI_ISL_440430|202 ···················· ···················· ····················· ···················· ···················· ····················
B 5553 hCoV-19/England/CAMB-74F64/2020|EPI_ISL_440431|202 ···················· ···················· ····················· ···················· ···················· ····················
B 5554 hCoV-19/England/CAMB-7514F/2020|EPI_ISL_440432|202 ···················· ···················· ····················· ···················· ···················· ····················
B 5555 hCoV-19/England/CAMB-750D9/2020|EPI_ISL_440433|202 ···················· ···················· ····················· ···················· ···················· ····················
B 5556 hCoV-19/England/CAMB-75200/2020|EPI_ISL_440434|202 ···················· ···················· ····················· ···················· ···················· ····················
B 5557 hCoV-19/England/CAMB-750F7/2020|EPI_ISL_440436|202 ···················· ···················· ····················· ···················· ···················· ····················
B 5558 hCoV-19/England/CAMB-75358/2020|EPI_ISL_440437|202 ···················· ···················· ····················· ···················· ···················· ····················
B 5559 hCoV-19/England/CAMB-75015/2020|EPI_ISL_440438|202 ···················· ···················· ····················· ···················· ···················· ····················
B 5560 hCoV-19/England/CAMB-84E19/2020|EPI_ISL_440439|202 ···················· ···················· ····················· ···················· ···················· ····················
B 5561 hCoV-19/England/CAMB-84EDD/2020|EPI_ISL_440445|202 ···················· ···················· ····················· ···················· ···················· ····················
B 5562 hCoV-19/England/CAMB-84F70/2020|EPI_ISL_440451|202 ···················· ···················· ····················· ···················· ···················· ····················
B 5563 hCoV-19/England/CAMB-84D94/2020|EPI_ISL_440460|202 ···················· ···················· ····················· ···················· ···················· ····················
B 5564 hCoV-19/England/CAMB-84B21/2020|EPI_ISL_440480|202 ···················· ···················· ····················· ···················· ···················· ····················
B 5565 hCoV-19/England/CAMB-84769/2020|EPI_ISL_440483|202 ···················· ···················· ····················· ···················· ···················· ····················
B 5566 hCoV-19/England/CAMB-8475A/2020|EPI_ISL_440486|202 ···················· ···················· ····················· ···················· ···················· ····················
B 5567 hCoV-19/England/CAMB-84ACA/2020|EPI_ISL_440487|202 ···················· ···················· ····················· ···················· ···················· ····················
B 5568 hCoV-19/England/CAMB-84BA9/2020|EPI_ISL_440489|202 ···················· ···················· ····················· ···················· ···················· ····················
B 5569 hCoV-19/England/CAMB-84918/2020|EPI_ISL_440491|202 ···················· ···················· ····················· ···················· ···················· ····················
B 5570 hCoV-19/England/CAMB-846B7/2020|EPI_ISL_440492|202 ···················· ···················· ····················· ···················· ···················· ····················
B 5571 hCoV-19/England/CAMB-847C3/2020|EPI_ISL_440494|202 ···················· ···················· ····················· ···················· ···················· ····················
B 5572 hCoV-19/England/CAMB-7D4B7/2020|EPI_ISL_441740|202 ···················· ···················· ····················· ···················· ···················· ····················
B 5573 hCoV-19/England/CAMB-7DB85/2020|EPI_ISL_441741|202 ···················· ···················· ····················· ···················· ···················· ····················
B 5574 hCoV-19/England/CAMB-7DB67/2020|EPI_ISL_441742|202 ···················· ···················· ····················· ···················· ···················· ····················
B 5575 hCoV-19/England/CAMB-7D44E/2020|EPI_ISL_441743|202 ···················· ···················· ····················· ···················· ···················· ····················
B 5576 hCoV-19/England/CAMB-7D727/2020|EPI_ISL_441744|202 ···················· ···················· ····················· ···················· ···················· ····················
B 5577 hCoV-19/England/CAMB-7DAB5/2020|EPI_ISL_441745|202 ···················· ···················· ····················· ···················· ···················· ····················
B 5578 hCoV-19/England/CAMB-7DB94/2020|EPI_ISL_441746|202 ···················· ···················· ····················· ···················· ···················· ····················
B 5579 hCoV-19/Northern Ireland/NIRE-FA9C2/2020|EPI_ISL_4 ···················· ···················· ····················· ···················· ···················· ····················
B 5580 hCoV-19/England/CAMB-7D657/2020|EPI_ISL_441748|202 ···················· ···················· ····················· ···················· ···················· ····················
```

| | | | | | | | | |
|---|---|---|---|---|---|---|---|---|
| B | 5581 | hCoV-19/England/CAMB-7D94F/2020\|EPI_ISL_441749\|202 | ···················· | ···················· | ···················· | ···················· | ···················· | ···················· |
| B | 5582 | hCoV-19/England/CAMB-7D99A/2020\|EPI_ISL_441750\|202 | ···················· | ···················· | ···················· | ···················· | ···················· | ···················· |
| B | 5583 | hCoV-19/Northern Ireland/NIRE-FA94A/2020\|EPI_ISL_4 | ···················· | ···················· | ···················· | ···················· | ···················· | ···················· |
| B | 5584 | hCoV-19/England/CAMB-7DCDD/2020\|EPI_ISL_441752\|202 | ···················· | ···················· | ···················· | ···················· | ···················· | ···················· |
| B | 5585 | hCoV-19/England/CAMB-7D9A9/2020\|EPI_ISL_441754\|202 | ···················· | ···················· | ···················· | ···················· | ···················· | ···················· |
| B | 5586 | hCoV-19/England/CAMB-7D5D2/2020\|EPI_ISL_441755\|202 | ···················· | ···················· | ···················· | ···················· | ···················· | ···················· |
| B | 5587 | hCoV-19/England/CAMB-7D8E8/2020\|EPI_ISL_441758\|202 | ···················· | ···················· | ···················· | ···················· | ···················· | ···················· |
| B | 5588 | hCoV-19/England/CAMB-7D763/2020\|EPI_ISL_441759\|202 | ···················· | ···················· | ···················· | ···················· | ···················· | ···················· |
| B | 5589 | hCoV-19/England/CAMB-7DC19/2020\|EPI_ISL_441760\|202 | ···················· | ···················· | ···················· | ···················· | ···················· | ···················· |
| B | 5590 | hCoV-19/England/CAMB-7D639/2020\|EPI_ISL_441762\|202 | ···················· | ···················· | ···················· | ···················· | ···················· | ···················· |
| B | 5591 | hCoV-19/England/CAMB-7D4C6/2020\|EPI_ISL_441764\|202 | ···················· | ···················· | ···················· | ···················· | ···················· | ···················· |
| B | 5592 | hCoV-19/Northern Ireland/NIRE-FAC8D/2020\|EPI_ISL_4 | ···················· | ···················· | ···················· | ···················· | ···················· | ···················· |
| B | 5593 | hCoV-19/Northern Ireland/NIRE-FA652/2020\|EPI_ISL_4 | ···················· | ···················· | ···················· | ···················· | ···················· | ···················· |
| B | 5594 | hCoV-19/England/CAMB-7DB1C/2020\|EPI_ISL_441767\|202 | ···················· | ···················· | ···················· | ···················· | ···················· | ···················· |
| B | 5595 | hCoV-19/England/CAMB-7DB0D/2020\|EPI_ISL_441768\|202 | ···················· | ···················· | ···················· | ···················· | ···················· | ···················· |
| B | 5596 | hCoV-19/England/CAMB-7D693/2020\|EPI_ISL_441769\|202 | ···················· | ···················· | ···················· | ···················· | ···················· | ···················· |
| B | 5597 | hCoV-19/Northern Ireland/NIRE-FAC6F/2020\|EPI_ISL_4 | ···················· | ···················· | ···················· | ···················· | ···················· | ···················· |
| B | 5598 | hCoV-19/Northern Ireland/NIRE-FA91D/2020\|EPI_ISL_4 | ···················· | ···················· | ···················· | ···················· | ···················· | ···················· |
| B | 5599 | hCoV-19/England/CAMB-7D7AF/2020\|EPI_ISL_441772\|202 | ···················· | ···················· | ···················· | ···················· | ···················· | ···················· |
| B | 5600 | hCoV-19/England/CAMB-7D97C/2020\|EPI_ISL_441773\|202 | ···················· | ···················· | ···················· | ···················· | ···················· | ···················· |
| B | 5601 | hCoV-19/England/CAMB-7DAF1/2020\|EPI_ISL_441774\|202 | ···················· | ···················· | ···················· | ···················· | ···················· | ···················· |
| B | 5602 | hCoV-19/Northern Ireland/NIRE-FA670/2020\|EPI_ISL_4 | ···················· | ···················· | ···················· | ···················· | ···················· | ···················· |
| B | 5603 | hCoV-19/England/CAMB-7DA5B/2020\|EPI_ISL_441777\|202 | ···················· | ···················· | ···················· | ···················· | ···················· | ···················· |
| B | 5604 | hCoV-19/England/CAMB-7DA00/2020\|EPI_ISL_441778\|202 | ···················· | ···················· | ···················· | ···················· | ···················· | ···················· |
| B | 5605 | hCoV-19/England/CAMB-7DBFE/2020\|EPI_ISL_441779\|202 | ···················· | ···················· | ···················· | ···················· | ···················· | ···················· |
| B | 5606 | hCoV-19/England/CAMB-7DAC4/2020\|EPI_ISL_441780\|202 | ···················· | ···················· | ···················· | ···················· | ···················· | ···················· |
| B | 5607 | hCoV-19/Northern Ireland/NIRE-FA9B3/2020\|EPI_ISL_4 | ···················· | ···················· | ···················· | ···················· | ···················· | ···················· |
| B | 5608 | hCoV-19/England/CAMB-7D587/2020\|EPI_ISL_441782\|202 | ···················· | ···················· | ···················· | ···················· | ···················· | ···················· |
| B | 5609 | hCoV-19/England/CAMB-7A60F/2020\|EPI_ISL_441784\|202 | ···················· | ···················· | ···················· | ···················· | ···················· | ···················· |
| B | 5610 | hCoV-19/England/CAMB-7903F/2020\|EPI_ISL_441785\|202 | ···················· | ···················· | ···················· | ···················· | ···················· | ···················· |
| B | 5611 | hCoV-19/England/CAMB-78AF6/2020\|EPI_ISL_441786\|202 | ···················· | ···················· | ···················· | ···················· | ···················· | ···················· |
| B | 5612 | hCoV-19/England/CAMB-7A3DB/2020\|EPI_ISL_441787\|202 | ···················· | ··········T········· | ···················· | ···················· | ···················· | ···················· |
| B | 5613 | hCoV-19/England/CAMB-79873/2020\|EPI_ISL_441788\|202 | ···················· | ···················· | ···················· | ···················· | ···················· | ···················· |
| B | 5614 | hCoV-19/England/CAMB-77FAD/2020\|EPI_ISL_441789\|202 | ···················· | ···················· | ···················· | ···················· | ···················· | ···················· |
| B | 5615 | hCoV-19/England/CAMB-7A2B0/2020\|EPI_ISL_441790\|202 | ···················· | ···················· | ···················· | ···················· | ···················· | ···················· |
| B | 5616 | hCoV-19/England/CAMB-790C6/2020\|EPI_ISL_441791\|202 | ···················· | ···················· | ···················· | ···················· | ···················· | ···················· |
| B | 5617 | hCoV-19/England/CAMB-79503/2020\|EPI_ISL_441792\|202 | ···················· | ···················· | ···················· | ···················· | ···················· | ···················· |
| B | 5618 | hCoV-19/England/CAMB-7922A/2020\|EPI_ISL_441793\|202 | ···················· | ···················· | ···················· | ···················· | ···················· | ···················· |
| B | 5619 | hCoV-19/Chile/LI-ISPCH-9/2020\|EPI_ISL_445300\|2020- | ···················· | ···················· | ···················· | ···················· | ···················· | ···················· |
| B | 5620 | hCoV-19/Chile/RM-ISPCH-23/2020\|EPI_ISL_445302\|2020 | ···················· | ···················· | ···················· | ···················· | ···················· | ···················· |
| B | 5621 | hCoV-19/Chile/RM-ISPCH-24/2020\|EPI_ISL_445303\|2020 | ···················· | ···················· | ···················· | ···················· | ···················· | ···················· |
| B | 5622 | hCoV-19/Wales/PHWC-32032/2020\|EPI_ISL_446600\|2020- | ···················· | ···················· | ···················· | ···················· | ···················· | ···················· |
| B | 5623 | hCoV-19/Wales/PHWC-32050/2020\|EPI_ISL_446601\|2020- | ···················· | ···················· | ···················· | ···················· | ···················· | ···················· |
| B | 5624 | hCoV-19/Wales/PHWC-3213F/2020\|EPI_ISL_446609\|2020- | ···················· | ···················· | ···················· | ···················· | ···················· | ···················· |
| B | 5625 | hCoV-19/Wales/PHWC-3218A/2020\|EPI_ISL_446612\|2020- | ···················· | ···················· | ···················· | ···················· | ···················· | ···················· |
| B | 5626 | hCoV-19/Wales/PHWC-321A8/2020\|EPI_ISL_446614\|2020- | ···················· | ···················· | ···················· | ···················· | ···················· | ···················· |
| B | 5627 | hCoV-19/Wales/PHWC-32296/2020\|EPI_ISL_446626\|2020- | ···················· | ···················· | ···················· | ···················· | ···················· | ···················· |
| B | 5628 | hCoV-19/Wales/PHWC-322B4/2020\|EPI_ISL_446628\|2020- | ···················· | ···················· | ···················· | ···················· | ···················· | ···················· |
| B | 5629 | hCoV-19/Wales/PHWC-322C3/2020\|EPI_ISL_446629\|2020- | ···················· | ···················· | ···················· | ···················· | ···················· | ···················· |
| B | 5630 | hCoV-19/Wales/PHWC-3232A/2020\|EPI_ISL_446634\|2020- | ···················· | ···················· | ···················· | ···················· | ···················· | ···················· |
| B | 5631 | hCoV-19/USA/FL-UF-4/2020\|EPI_ISL_524433\|2020-03-27 | ···················· | ···················· | ···················· | ···················· | ···················· | ···············A···· |
| B | 5632 | hCoV-19/USA/FL-UF-5/2020\|EPI_ISL_524434\|2020-03-27 | ···················· | ···················· | ···················· | ···················· | ···················· | ···················· |
| B | 5633 | hCoV-19/Scotland/EDB582/2020\|EPI_ISL_433204\|2020-0 | ···················· | ···················· | ···················· | ···················· | ···················· | ···················· |
| B | 5634 | hCoV-19/Scotland/EDB587/2020\|EPI_ISL_433205\|2020-0 | ···················· | ···················· | ···················· | ···················· | ···················· | ···················· |
| B | 5635 | hCoV-19/Scotland/EDB588/2020\|EPI_ISL_433206\|2020-0 | ···················· | ···················· | ···················· | ···················· | ···················· | ···················· |
| B | 5636 | hCoV-19/Scotland/EDB589/2020\|EPI_ISL_433207\|2020-0 | ···················· | ···················· | ···················· | ···················· | ··············N····· | NN·················· |
| B | 5639 | hCoV-19/Scotland/EDB598/2020\|EPI_ISL_433211\|2020-0 | ···················· | ···················· | ···················· | ···················· | ···················· | ···················· |
| B | 5641 | hCoV-19/Scotland/EDB607/2020\|EPI_ISL_433214\|2020-0 | ···················· | ···················· | ···················· | ···················· | ···················· | ···················· |
| B | 5642 | hCoV-19/Scotland/EDB612/2020\|EPI_ISL_433215\|2020-0 | ·T·················· | ···················· | ···················· | ···················· | ···················· | ···················· |
| B | 5643 | hCoV-19/Scotland/EDB613/2020\|EPI_ISL_433216\|2020-0 | ···················· | ·············A······ | ···················· | ···················· | ···················· | ···················· |
| B | 5644 | hCoV-19/Scotland/EDB620/2020\|EPI_ISL_433217\|2020-0 | ···················· | ···················· | ···················· | ···················· | ···················· | ···················· |
| B | 5645 | hCoV-19/Scotland/EDB625/2020\|EPI_ISL_433218\|2020-0 | ···················· | ···················· | ···················· | ···················· | ···················· | ···················· |
| B | 5648 | hCoV-19/Scotland/EDB682/2020\|EPI_ISL_433222\|2020-0 | ···················· | ·A·················· | ···················· | ···················· | ···················· | ···················· |
| B | 5649 | hCoV-19/Scotland/EDB689/2020\|EPI_ISL_433223\|2020-0 | ···················· | ·A·················· | ···················· | ···················· | ···················· | ···················· |
| B | 5650 | hCoV-19/Scotland/EDB694/2020\|EPI_ISL_433224\|2020-0 | ···················· | ···················· | ···················· | ···················· | ···················· | ···················· |
| B | 5651 | hCoV-19/Scotland/EDB695/2020\|EPI_ISL_433225\|2020-0 | ···················· | ···················· | ···················· | ···················· | ···················· | ···················· |
| B | 5652 | hCoV-19/Scotland/EDB703/2020\|EPI_ISL_433226\|2020-0 | ···················· | ···················· | ···················· | ···················· | ···················· | ···················· |
| B | 5653 | hCoV-19/Scotland/EDB707/2020\|EPI_ISL_433227\|2020-0 | ···················· | ···················· | ···················· | ···················· | ···················· | ···················· |
| B | 5654 | hCoV-19/Scotland/EDB721/2020\|EPI_ISL_433228\|2020-0 | ···················· | ···················· | ···················· | ···················· | ···················· | ···················· |
| B | 5655 | hCoV-19/Scotland/EDB726/2020\|EPI_ISL_433229\|2020-0 | ···················· | ···················· | ···················· | ···················· | ···················· | ···················· |
| B | 5656 | hCoV-19/Scotland/EDB734/2020\|EPI_ISL_433233\|2020-0 | ···················· | ···················· | ···················· | ···················· | ···················· | ···················· |
| B | 5657 | hCoV-19/Scotland/EDB735/2020\|EPI_ISL_433234\|2020-0 | ···················· | ···················· | ···················· | ···················· | ···················· | ···················· |
| B | 5658 | hCoV-19/Scotland/EDB753/2020\|EPI_ISL_433236\|2020-0 | ···················· | ·A·················· | ···················· | ···················· | ···················· | ···················· |
| B | 5659 | hCoV-19/Scotland/EDB764/2020\|EPI_ISL_433237\|2020-0 | ···················· | ···················· | ···················· | ···················· | ···················· | ···················· |
| B | 5660 | hCoV-19/Scotland/EDB766/2020\|EPI_ISL_433238\|2020-0 | ···················· | ···················· | ···················· | ···················· | ···················· | ···················· |
| B | 5661 | hCoV-19/Scotland/EDB773/2020\|EPI_ISL_433240\|2020-0 | ···················· | ···················· | ···················· | ···················· | ···················· | ···················· |
| B | 5662 | hCoV-19/Scotland/EDB774/2020\|EPI_ISL_433241\|2020-0 | ···················· | ···················A | ···················· | ···················· | ···················· | ···················· 1 |
| B | 5663 | hCoV-19/Scotland/EDB781/2020\|EPI_ISL_433242\|2020-0 | ···················· | ·A·················· | ···················· | ···················· | ···················· | ···················· |

| | | | | | | | |
|---|---|---|---|---|---|---|---|
| B | 5664 hCoV-19/Scotland/EDB787/2020\|EPI_ISL_433243\|2020-0 | ···················· | ···················· | ····················· | ···················· | ···················· | ···················· |
| B | 5665 hCoV-19/Scotland/EDB788/2020\|EPI_ISL_433244\|2020-0 | ···················· | ···················· | ····················· | ···················· | ···················· | ···················· |
| B | 5666 hCoV-19/Scotland/EDB789/2020\|EPI_ISL_433245\|2020-0 | ···················· | ···················· | ····················· | ···················· | ···················· | ···················· |
| B | 5667 hCoV-19/Scotland/EDB795/2020\|EPI_ISL_433246\|2020-0 | ···················· | ···················· | ····················· | ···················· | ···················· | ···················· |
| B | 5668 hCoV-19/Scotland/EDB799/2020\|EPI_ISL_433247\|2020-0 | ···················· | ···················· | ····················· | ···················· | ···················· | ···················· |
| B | 5669 hCoV-19/Scotland/EDB801/2020\|EPI_ISL_433248\|2020-0 | ···················· | ···················· | ····················· | ···················· | ···················· | ···················· |
| B | 5670 hCoV-19/Scotland/EDB807/2020\|EPI_ISL_433249\|2020-0 | ···················· | ···················· | ····················· | ···················· | ···················· | ···················· |
| B | 5671 hCoV-19/Scotland/EDB821/2020\|EPI_ISL_433250\|2020-0 | ···················· | ···················· | ····················· | ···················· | ···················· | ···················· |
| B | 5672 hCoV-19/Scotland/EDB838/2020\|EPI_ISL_433253\|2020-0 | ···················· | ···················· | ····················· | ···················· | ···················· | ···················· |
| B | 5673 hCoV-19/Scotland/EDB842/2020\|EPI_ISL_433254\|2020-0 | ···················· | ···················· | ····················· | ···················· | ···················· | ···················· |
| B | 5674 hCoV-19/Scotland/EDB848/2020\|EPI_ISL_433255\|2020-0 | ···················· | ·A·················· | ····················· | ···················· | ···················· | ···················· |
| B | 5675 hCoV-19/Scotland/EDB852/2020\|EPI_ISL_433256\|2020-0 | ···················· | ···················· | ····················· | ···················· | ···················· | ···················· |
| B | 5676 hCoV-19/Scotland/EDB860/2020\|EPI_ISL_433257\|2020-0 | ···················· | ·A·················· | ····················· | ···················· | ···················· | ···················· |
| B | 5677 hCoV-19/Scotland/EDB888/2020\|EPI_ISL_433266\|2020-0 | ···················· | ···················· | ····················· | ···················· | ···················· | ···················· |
| B | 5678 hCoV-19/Scotland/EDB889/2020\|EPI_ISL_433267\|2020-0 | ···················· | ···················· | ····················· | ···················· | ···················· | ···················· |
| B | 5679 hCoV-19/Scotland/CVR1978/2020\|EPI_ISL_433270\|2020- | ···················· | ···················· | ····················· | ···················· | ···················· | ···················· |
| B | 5680 hCoV-19/Scotland/CVR225/2020\|EPI_ISL_433274\|2020-0 | ···················· | ···················· | ····················· | ···················· | ···················· | ···················· |
| B | 5681 hCoV-19/Scotland/CVR248/2020\|EPI_ISL_433276\|2020-0 | ···················· | ·A·················· | ····················· | ···················· | ···················· | ···················· |
| B | 5682 hCoV-19/Scotland/CVR261/2020\|EPI_ISL_433277\|2020-0 | ···················· | ···················· | ····················· | ···················· | ···················· | ···················· |
| B | 5683 hCoV-19/Scotland/CVR273/2020\|EPI_ISL_433278\|2020-0 | ···················· | ···················· | ····················· | ···················· | ···················· | ···················· |
| B | 5684 hCoV-19/Scotland/CVR278/2020\|EPI_ISL_433279\|2020-0 | ···················· | ···················· | ····················· | ···················· | ···················· | ···················· |
| B | 5685 hCoV-19/Scotland/CVR282/2020\|EPI_ISL_433280\|2020-0 | ···················· | ···················· | ····················· | ···················· | ···················· | ···················· |
| B | 5686 hCoV-19/Scotland/CVR284/2020\|EPI_ISL_433281\|2020-0 | ···················· | ·A·················· | ····················· | ···················· | ···················· | ···················· |
| B | 5687 hCoV-19/Scotland/CVR304/2020\|EPI_ISL_433282\|2020-0 | ···················· | ···················· | ····················· | ···················· | ···················· | ···················· |
| B | 5688 hCoV-19/Scotland/CVR309/2020\|EPI_ISL_433283\|2020-0 | ···················· | ·A·················· | ····················· | ···················· | ···················· | ···················· |
| B | 5689 hCoV-19/Scotland/CVR338/2020\|EPI_ISL_433284\|2020-0 | ···················· | ···················· | ····················· | ···················· | ···················· | ···················· |
| B | 5690 hCoV-19/Scotland/CVR364/2020\|EPI_ISL_433285\|2020-0 | ···················· | ···················· | ····················· | ···················· | ···················· | ···················· |
| B | 5691 hCoV-19/Scotland/CVR367/2020\|EPI_ISL_433286\|2020-0 | ···················· | ···················· | ····················· | ···················· | ···················· | ···················· |
| B | 5692 hCoV-19/Scotland/CVR375/2020\|EPI_ISL_433287\|2020-0 | ···················· | ···················· | ····················· | ···················· | ···················· | ···················· |
| B | 5693 hCoV-19/Scotland/CVR392/2020\|EPI_ISL_433288\|2020-0 | ···················· | ·A·················· | ····················· | ···················· | ···················· | ···················· |
| B | 5694 hCoV-19/Scotland/CVR397/2020\|EPI_ISL_433289\|2020-0 | ···················· | ···················· | ····················· | ···················· | ···················· | ···················· |
| B | 5695 hCoV-19/Scotland/CVR425/2020\|EPI_ISL_433291\|2020-0 | ···················· | ·A·················· | ····················· | ···················· | ···················· | ···················· |
| B | 5696 hCoV-19/Scotland/CVR436/2020\|EPI_ISL_433293\|2020-0 | ···················· | ···················· | ····················· | ···················· | ···················· | ···················· |
| B | 5697 hCoV-19/Scotland/CVR438/2020\|EPI_ISL_433294\|2020-0 | ···················· | ·A·················· | ····················· | ···················· | ···················· | ···················· |
| B | 5698 hCoV-19/Scotland/CVR450/2020\|EPI_ISL_433295\|2020-0 | ···················· | ···················· | ····················· | ···················· | ···················· | ···················· |
| B | 5699 hCoV-19/Scotland/CVR458/2020\|EPI_ISL_433298\|2020-0 | ···················· | ···················· | ····················· | ···················· | ···················· | ···················· |
| B | 5700 hCoV-19/Scotland/CVR464/2020\|EPI_ISL_433299\|2020-0 | ···················· | ···················· | ····················· | ···················· | ···················· | ···················· |
| B | 5701 hCoV-19/Costa Rica/02/2020\|EPI_ISL_434535\|2020-03- | ···················· | ···················· | ····················· | ···················· | ···················· | ···················· |
| B | 5702 hCoV-19/Costa Rica/05/2020\|EPI_ISL_434538\|2020-03- | ···················· | ···················· | ····················· | ···················· | ···················· | ···················· |
| B | 5703 hCoV-19/Costa Rica/06/2020\|EPI_ISL_434539\|2020-03- | ···················· | ···················· | ····················· | ···················· | ···················· | ···················· |
| B | 5704 hCoV-19/Puerto Rico/CDC-S1/2020\|EPI_ISL_434541\|202 | ···················· | ···················· | ····················· | ···················· | ···················· | ···················· |
| B | 5705 hCoV-19/Puerto Rico/CDC-S3/2020\|EPI_ISL_434543\|202 | ···················· | ···················· | ····················· | ···················· | ···················· | ···················· |
| B | 5706 hCoV-19/Puerto Rico/CDC-S5/2020\|EPI_ISL_434545\|202 | ···················· | ···················· | ····················· | ···················· | ···················· | ···················· |
| B | 5707 hCoV-19/Puerto Rico/CDC-S6/2020\|EPI_ISL_434546\|202 | ···················· | ···················· | ····················· | ···················· | ···················· | ···················· |
| B | 5708 hCoV-19/Puerto Rico/CDC-S7/2020\|EPI_ISL_434547\|202 | ···················· | ···················· | ····················· | ···················· | ···················· | ···················· |
| B | 5709 hCoV-19/Puerto Rico/CDC-S8/2020\|EPI_ISL_434548\|202 | ···················· | ···················· | ····················· | ···················· | ···················· | ···················· |
| B | 5710 hCoV-19/Puerto Rico/CDC-S9/2020\|EPI_ISL_434549\|202 | ···················· | ···················· | ····················· | ···················· | ···················· | ···················· |
| B | 5711 hCoV-19/Puerto Rico/CDC-S11/2020\|EPI_ISL_434551\|20 | ···················· | ···················· | ····················· | ···················· | ···················· | ···················· |
| B | 5712 hCoV-19/Philippines/PGC03/2020\|EPI_ISL_434555\|2020 | ···················· | ···················· | ····················· | ···················· | ···················· | ···················· |
| B | 5713 hCoV-19/Philippines/PGC06/2020\|EPI_ISL_434558\|2020 | ···················· | ···················· | ····················· | ···················· | ···················· | ···················· |
| B | 5714 hCoV-19/Czech Republic/2308/2020\|EPI_ISL_434572\|20 | ···················· | ···················· | ····················· | ···················· | ···················· | ···················· |
| B | 5715 hCoV-19/USA/MD-HP00036/2020\|EPI_ISL_434586\|2020-03 | ···················· | ···················· | ····················· | ···················· | ···················· | ···················· |
| B | 5716 hCoV-19/USA/MD-HP00037/2020\|EPI_ISL_434587\|2020-03 | ···················· | ···················· | ····················· | ···················· | ···················· | ···················· |
| B | 5717 hCoV-19/USA/MD-HP00038/2020\|EPI_ISL_434588\|2020-03 | ···················· | ···················· | ····················· | ···················· | ···················· | ···················· |
| B | 5718 hCoV-19/USA/VA-DCLS-0100/2020\|EPI_ISL_434594\|2020- | ···················· | ···················· | ····················· | ···················· | ···················· | ···················· |
| B | 5719 hCoV-19/USA/VA-DCLS-0105/2020\|EPI_ISL_434597\|2020- | ···················· | ···················· | ····················· | ···················· | ···················· | ···················· |
| B | 5720 hCoV-19/USA/VA-DCLS-0106/2020\|EPI_ISL_434598\|2020- | ···················· | ···················· | ····················· | ···················· | ···················· | ···················· |
| B | 5721 hCoV-19/USA/VA-DCLS-0107/2020\|EPI_ISL_434599\|2020- | ···················· | ···················· | ····················· | ···················· | ···················· | ···················· |
| B | 5722 hCoV-19/USA/WA-S597/2020\|EPI_ISL_438150\|2020-04-13 | ···················· | ···················· | ····················· | ···················· | ···················· | ···················· |
| B | 5723 hCoV-19/USA/WA-S598/2020\|EPI_ISL_438151\|2020-04-13 | ···················· | ···················· | ····················· | ···················· | ···················· | ···················· |
| B | 5724 hCoV-19/USA/WA-S599/2020\|EPI_ISL_438152\|2020-03-26 | ···················· | ···················· | ····················· | ···················· | ···················· | ···················· |
| B | 5725 hCoV-19/USA/WA-S600/2020\|EPI_ISL_438153\|2020-04-15 | ···················· | ···················· | ····················· | ···················· | ···················· | ···················· |
| B | 5726 hCoV-19/USA/WA-S601/2020\|EPI_ISL_438154\|2020-03-20 | ···················· | ···················· | ····················· | ···················· | ···················· | ···················· |
| B | 5727 hCoV-19/USA/WA-S602/2020\|EPI_ISL_438155\|2020-03-23 | ···················· | ···················· | ····················· | ···················· | ···················· | ···················· |
| B | 5728 hCoV-19/USA/WA-S603/2020\|EPI_ISL_438156\|2020-04-15 | ···················· | ···················· | ····················· | ···················· | ···················· | ···················· |
| B | 5729 hCoV-19/USA/WA-S604/2020\|EPI_ISL_438157\|2020-04-19 | ···················· | ···················· | ····················· | ···················· | ···················· | ···················· |
| B | 5730 hCoV-19/USA/WA-S605/2020\|EPI_ISL_438158\|2020-04-18 | ···················· | ···················· | ····················· | ···················· | ···················· | ···················· |
| B | 5731 hCoV-19/USA/WA-S606/2020\|EPI_ISL_438159\|2020-04-20 | ···················· | ···················· | ····················· | ···················· | ···················· | ···················· |
| B | 5732 hCoV-19/England/CAMB-76E29/2020\|EPI_ISL_439400\|202 | ···················· | ···················· | ····················· | ···················· | ···················· | ···················· |
| B | 5733 hCoV-19/England/CAMB-77A51/2020\|EPI_ISL_439401\|202 | ···················· | ···················· | ····················· | ···················· | ···················· | ···················· |
| B | 5734 hCoV-19/England/CAMB-77189/2020\|EPI_ISL_439402\|202 | ···················· | ···················· | ····················· | ···················· | ···················· | ···················· |
| B | 5735 hCoV-19/England/CAMB-76D4A/2020\|EPI_ISL_439403\|202 | ···················· | ···················· | ····················· | ···················· | ···················· | ···················· |
| B | 5736 hCoV-19/England/CAMB-777A5/2020\|EPI_ISL_439404\|202 | ···················· | ···················· | ····················· | ···················· | ···················· | ···················· |
| B | 5737 hCoV-19/England/CAMB-77866/2020\|EPI_ISL_439405\|202 | ···················· | ···················· | ····················· | ···················· | ···················· | ···················· |
| B | 5738 hCoV-19/England/CAMB-7758D/2020\|EPI_ISL_439406\|202 | ···················· | ···················· | ····················· | ···················· | ···················· | ···················· |
| B | 5739 hCoV-19/England/CAMB-76050/2020\|EPI_ISL_439407\|202 | ···················· | ···················· | ····················· | ···················· | ···················· | ···················· |
| B | 5740 hCoV-19/England/CAMB-764DC/2020\|EPI_ISL_439408\|202 | ···················· | ···················· | ····················· | ···················· | ···················· | ···················· |
| B | 5741 hCoV-19/England/CAMB-77972/2020\|EPI_ISL_439409\|202 | ···················· | ···················· | ····················· | ···················· | ···················· | ···················· |

| | | | | | | | | |
|---|---|---|---|---|---|---|---|---|
| B | 5742 | hCoV-19/England/CAMB-7631B/2020\|EPI_ISL_439410\|202 | ···················· | ···················· | ···················· | ···················· | ···················· | ···················· |
| B | 5743 | hCoV-19/England/CAMB-76630/2020\|EPI_ISL_439411\|202 | ···················· | ···················· | ···················· | ···················· | ···················· | ···················· |
| B | 5744 | hCoV-19/England/CAMB-7772D/2020\|EPI_ISL_439414\|202 | ···················· | ···················· | ···················· | ···················· | ···················· | ···················· |
| B | 5745 | hCoV-19/England/CAMB-76E47/2020\|EPI_ISL_439415\|202 | ···················· | ···················· | ···················· | ···················· | ···················· | ···················· |
| B | 5746 | hCoV-19/England/CAMB-774F9/2020\|EPI_ISL_439416\|202 | ···················· | ···················· | ···················· | ···················· | ···················· | ···················· |
| B | 5747 | hCoV-19/England/CAMB-7715C/2020\|EPI_ISL_439417\|202 | ···················· | ···················· | ···················· | ···················· | ···················· | ···················· |
| B | 5748 | hCoV-19/England/CAMB-778EE/2020\|EPI_ISL_439418\|202 | ···················· | ···················· | ···················· | ···················· | ···················· | ···················· |
| B | 5749 | hCoV-19/England/CAMB-76603/2020\|EPI_ISL_439419\|202 | ···················· | ···················· | ···················· | ···················· | ···················· | ···················· |
| B | 5750 | hCoV-19/England/CAMB-772D1/2020\|EPI_ISL_439420\|202 | ···················· | ···················· | ···················· | ···················· | ···················· | ···················· |
| B | 5751 | hCoV-19/England/CAMB-7775A/2020\|EPI_ISL_439421\|202 | ···················· | ···················· | ···················· | ···················· | ···················· | ···················· |
| B | 5752 | hCoV-19/England/CAMB-7771E/2020\|EPI_ISL_439422\|202 | ···················· | ···················· | ···················· | ···················· | ···················· | ···················· |
| B | 5753 | hCoV-19/England/CAMB-77602/2020\|EPI_ISL_439423\|202 | ···················· | ···················· | ···················· | ···················· | ···················· | ···················· |
| B | 5754 | hCoV-19/England/CAMB-7749F/2020\|EPI_ISL_439424\|202 | ···················· | ···················· | ···················· | ···················· | ···················· | ···················· |
| B | 5755 | hCoV-19/England/CAMB-7764E/2020\|EPI_ISL_439426\|202 | ···················· | ···················· | ···················· | ···················· | ···················· | ···················· |
| B | 5756 | hCoV-19/England/CAMB-772A4/2020\|EPI_ISL_439428\|202 | ···················· | ···················· | ···················· | ···················· | ···················· | ···················· |
| B | 5757 | hCoV-19/England/CAMB-77AF7/2020\|EPI_ISL_439429\|202 | ···················· | ···················· | ···················· | ···················· | ···················· | ···················· |
| B | 5758 | hCoV-19/England/CAMB-777E1/2020\|EPI_ISL_439481\|202 | ···················· | ···················· | ···················· | ···················· | ···················· | ···················· |
| B | 5759 | hCoV-19/England/CAMB-75B05/2020\|EPI_ISL_439482\|202 | ···················· | ···················· | ···················· | ···················· | ···················· | ···················· |
| B | 5760 | hCoV-19/England/CAMB-779EB/2020\|EPI_ISL_439484\|202 | ···················· | ···················· | ···················· | ···················· | ···················· | ···················· |
| B | 5761 | hCoV-19/England/CAMB-773DE/2020\|EPI_ISL_439485\|202 | ···················· | ···················· | ···················· | ···················· | ···················· | ···················· |
| B | 5762 | hCoV-19/England/CAMB-764BE/2020\|EPI_ISL_439486\|202 | ···················· | ···················· | ···················· | ···················· | ···················· | ···················· |
| B | 5763 | hCoV-19/England/CAMB-772E0/2020\|EPI_ISL_439487\|202 | ···················· | ···················· | ···················· | ···················· | ···················· | ···················· |
| B | 5764 | hCoV-19/England/CAMB-77875/2020\|EPI_ISL_439489\|202 | ···T················ | ···················· | ···················· | ···················· | ···················· | ···················· |
| B | 5765 | hCoV-19/England/CAMB-75130/2020\|EPI_ISL_440400\|202 | ···················· | ···················· | ···················· | ···················· | ···················· | ···················· |
| B | 5766 | hCoV-19/England/CAMB-75024/2020\|EPI_ISL_440401\|202 | ···················· | ···················· | ···················· | ···················· | ···················· | ···················· |
| B | 5767 | hCoV-19/England/CAMB-75033/2020\|EPI_ISL_440402\|202 | ···················· | ···················· | ···················· | ···················· | ···················· | ···················· |
| B | 5768 | hCoV-19/England/CAMB-7525B/2020\|EPI_ISL_440403\|202 | ···················· | ···················· | ···················· | ···················· | ···················· | ···················· |
| B | 5769 | hCoV-19/England/CAMB-751C7/2020\|EPI_ISL_440404\|202 | ···················· | ···················· | ···················· | ···················· | ···················· | ···················· |
| B | 5770 | hCoV-19/England/CAMB-7507F/2020\|EPI_ISL_440405\|202 | ···················· | ···················· | ···················· | ···················· | ···················· | ···················· |
| B | 5772 | hCoV-19/England/CAMB-750AC/2020\|EPI_ISL_440409\|202 | ···················· | ···················· | ···················· | ···················· | ···················· | ···················· |
| B | 5773 | hCoV-19/England/CAMB-84E37/2020\|EPI_ISL_440459\|202 | ···················· | ···················· | ···················· | ···················· | ···················· | ···················· |
| B | 5774 | hCoV-19/England/CAMB-84F9E/2020\|EPI_ISL_440464\|202 | ···················· | ···················· | ···················· | ···················· | ···················· | ···················· |
| B | 5775 | hCoV-19/England/CAMB-7DBA3/2020\|EPI_ISL_441700\|202 | ···················· | ···················· | ···················· | ···················· | ···················· | ···················· |
| B | 5776 | hCoV-19/England/CAMB-7D912/2020\|EPI_ISL_441701\|202 | ···················· | ···················· | ···················· | ···················· | ···················· | ···················· |
| B | 5777 | hCoV-19/England/CAMB-7D930/2020\|EPI_ISL_441702\|202 | ···················· | ···················· | ···················· | ···················· | ···················· | ···················· |
| B | 5778 | hCoV-19/England/CAMB-7DB76/2020\|EPI_ISL_441703\|202 | ···················· | ···················· | ···················· | ···················· | ···················· | ···················· |
| B | 5779 | hCoV-19/England/CAMB-7D8D9/2020\|EPI_ISL_441707\|202 | ···················· | ···················· | ···················· | ···················· | ···················· | ···················· |
| B | 5780 | hCoV-19/Northern Ireland/NIRE-FAA83/2020\|EPI_ISL_4 | ···················· | ···················· | ···················· | ···················· | ···················· | ···················· |
| B | 5781 | hCoV-19/England/CAMB-7D45D/2020\|EPI_ISL_441709\|202 | ···················· | ···················· | ···················· | ···················· | ···················· | ···················· |
| B | 5782 | hCoV-19/England/CAMB-7D745/2020\|EPI_ISL_441711\|202 | ···················· | ···················· | ···················· | ···················· | ···················· | ···················· |
| B | 5783 | hCoV-19/England/CAMB-7D842/2020\|EPI_ISL_441713\|202 | ···················· | ···················· | ···················· | ···················· | ···················· | ···················· |
| B | 5784 | hCoV-19/Northern Ireland/NIRE-FAACF/2020\|EPI_ISL_4 | ···················· | ···················· | ···················· | ···················· | ···················· | ···················· |
| B | 5785 | hCoV-19/England/CAMB-7DC91/2020\|EPI_ISL_441715\|202 | ···················· | ···················· | ···················· | ···················· | ···················· | ···················· |
| B | 5786 | hCoV-19/Northern Ireland/NIRE-FA616/2020\|EPI_ISL_4 | ···················· | ···················· | ···················· | ···················· | ···················· | ···················· |
| B | 5787 | hCoV-19/England/CAMB-7D4E4/2020\|EPI_ISL_441718\|202 | ···················· | ···················· | ···················· | ···················· | ···················· | ···················· |
| B | 5788 | hCoV-19/England/CAMB-7D675/2020\|EPI_ISL_441720\|202 | ···················· | ···················· | ···················· | ···················· | ···················· | ···················· |
| B | 5789 | hCoV-19/England/CAMB-7D52D/2020\|EPI_ISL_441721\|202 | ···················· | ···················· | ···················· | ···················· | ···················· | ···················· |
| B | 5790 | hCoV-19/England/CAMB-7D7EB/2020\|EPI_ISL_441723\|202 | ···················· | ···················· | ···················· | ···················· | ···················· | ···················· |
| B | 5791 | hCoV-19/England/CAMB-7DB58/2020\|EPI_ISL_441724\|202 | ···················· | ···················· | ···················· | ···················· | ···················· | ···················· |
| B | 5792 | hCoV-19/England/CAMB-7DA79/2020\|EPI_ISL_441725\|202 | ···················· | ···················· | ···················· | ···················· | ···················· | ···················· |
| B | 5793 | hCoV-19/England/CAMB-7DB2B/2020\|EPI_ISL_441726\|202 | ···················· | ···················· | ···················· | ···················· | ···················· | ···················· |
| B | 5794 | hCoV-19/England/CAMB-7DB3A/2020\|EPI_ISL_441727\|202 | ···················· | ···················· | ···················· | ···················· | ···················· | ···················· |
| B | 5795 | hCoV-19/England/CAMB-7D54B/2020\|EPI_ISL_441728\|202 | ···················· | ···················· | ···················· | ···················· | ···················· | ···················· |
| B | 5796 | hCoV-19/England/CAMB-7D5F0/2020\|EPI_ISL_441729\|202 | ···················· | ···················· | ···················· | ···················· | ···················· | ···················· |
| B | 5797 | hCoV-19/Northern Ireland/NIRE-FAB08/2020\|EPI_ISL_4 | ···················· | ···················· | ···················· | ···················· | ···················· | ···················· |
| B | 5798 | hCoV-19/England/CAMB-7D48A/2020\|EPI_ISL_441731\|202 | ···················· | ···················· | ···················· | ···················· | ···················· | ···················· |
| B | 5799 | hCoV-19/England/CAMB-7D7CD/2020\|EPI_ISL_441732\|202 | ···················· | ···················· | ···················· | ···················· | ···················· | ···················· |
| B | 5800 | hCoV-19/Northern Ireland/NIRE-FA69E/2020\|EPI_ISL_4 | ···················· | ···················· | ···················· | ···················· | ···················· | ···················· |
| B | 5801 | hCoV-19/England/CAMB-7DB49/2020\|EPI_ISL_441734\|202 | ···················· | ···················· | ···················· | ···················· | ···················· | ···················· |
| B | 5802 | hCoV-19/England/CAMB-7D9C7/2020\|EPI_ISL_441735\|202 | ···················· | ···················· | ···················· | ···················· | ···················· | ···················· |
| B | 5803 | hCoV-19/England/CAMB-7D781/2020\|EPI_ISL_441736\|202 | ···················· | ···················· | ···················· | ···················· | ···················· | ···················· |
| B | 5804 | hCoV-19/Northern Ireland/NIRE-FAAED/2020\|EPI_ISL_4 | ···················· | ···················· | ···················· | ···················· | ···················· | ···················· |
| B | 5805 | hCoV-19/England/CAMB-7D6B1/2020\|EPI_ISL_441738\|202 | ···················· | ···················· | ···················· | ···················· | ···················· | ···················· |
| B | 5806 | hCoV-19/England/CAMB-7D806/2020\|EPI_ISL_441739\|202 | ···················· | ···················· | ···················· | ···················· | ···················· | ···················· |
| B | 5807 | hCoV-19/England/CAMB-77FCB/2020\|EPI_ISL_441795\|202 | ···················· | ···················· | ···················· | ···················· | ···················· | ···················· |
| B | 5808 | hCoV-19/England/CAMB-79819/2020\|EPI_ISL_441797\|202 | ···················· | ···················· | ···················· | ···················· | ···················· | ···················· |
| B | 5809 | hCoV-19/England/CAMB-78FD9/2020\|EPI_ISL_441798\|202 | ···················· | ···················· | ···················· | ···················· | ···················· | ···················· |
| B | 5810 | hCoV-19/Bangladesh/DNAS-CPH-471/2020\|EPI_ISL_44521 | ···················· | ···················· | ···················· | ···················· | ···················· | ···················· |
| B | 5811 | hCoV-19/Bangladesh/DNAS-CPH-427/2020\|EPI_ISL_44521 | ···················· | ···················· | ···················· | ···················· | ···················· | ···················· |
| B | 5812 | hCoV-19/Bangladesh/DNAS-CPH-466/2020\|EPI_ISL_44521 | ···················· | ···················· | ···················· | ···················· | ···················· | ···················· |
| B | 5813 | hCoV-19/Colombia/VAC-UdeV-01-2/2020\|EPI_ISL_445219 | ···················· | ···················· | ···················· | ···················· | ···················· | ···················· |
| B | 5814 | hCoV-19/Romania/278406/2020\|EPI_ISL_445220\|2020-04 | ···················· | ···················· | ···················· | ···················· | ···················· | ···················· |
| B | 5815 | hCoV-19/Sweden/20-05444/2020\|EPI_ISL_445221\|2020-0 | ···················· | ···················· | ···················· | ···················· | ···················· | ···················· |
| B | 5816 | hCoV-19/Sweden/20-05823/2020\|EPI_ISL_445222\|2020-0 | ···················· | ···················· | ···················· | ···················· | ···················· | ···················· |
| B | 5817 | hCoV-19/Sweden/20-06208/2020\|EPI_ISL_445223\|2020-0 | ···················· | ···················· | ···················· | ···················· | ···················· | ···················· |
| B | 5818 | hCoV-19/Sweden/20-06459/2020\|EPI_ISL_445224\|2020-0 | ···················· | ···················· | ···················· | ···················· | ···················· | ···················· |
| B | 5819 | hCoV-19/Sweden/20-06653/2020\|EPI_ISL_445225\|2020-0 | ···················· | ···················· | ···················· | ···················· | ···················· | ···················· |
| B | 5820 | hCoV-19/Sweden/20-06679/2020\|EPI_ISL_445226\|2020-0 | ···················· | ···················· | ···················· | ···················· | ···················· | ···················· |

| | | | | | | | | |
|---|---|---|---|---|---|---|---|---|
| B | 5821 | hCoV-19/Sweden/20-06805/2020\|EPI_ISL_445227\|2020-0 | .................... | .................... | ..................... | .................... | .................... | .................... |
| B | 5822 | hCoV-19/Sweden/20-06814/2020\|EPI_ISL_445228\|2020-0 | .................... | .................... | ..................... | .................... | .................... | .................... |
| B | 5823 | hCoV-19/Sweden/20-06817/2020\|EPI_ISL_445229\|2020-0 | .................... | .................... | ..................... | .................... | .................... | .................... |
| B | 5824 | hCoV-19/Sweden/20-07044/2020\|EPI_ISL_445230\|2020-0 | .................... | .................... | ..................... | .................... | .................... | .................... |
| B | 5825 | hCoV-19/Sweden/20-07237/2020\|EPI_ISL_445231\|2020-0 | .................... | .................... | ..................... | .................... | .................... | .................... |
| B | 5826 | hCoV-19/Sweden/20-07243/2020\|EPI_ISL_445232\|2020-0 | .................... | .................... | ..................... | .................... | .................... | .................... |
| B | 5827 | hCoV-19/Sweden/20-07452/2020\|EPI_ISL_445233\|2020-0 | .................... | .................... | ..................... | .................... | .................... | .................... |
| B | 5828 | hCoV-19/Sweden/20-07571/2020\|EPI_ISL_445234\|2020-0 | .................... | .................... | ..................... | .................... | .................... | .................... |
| B | 5829 | hCoV-19/Sweden/20-07586/2020\|EPI_ISL_445235\|2020-0 | .................... | .................... | ..................... | .................... | .................... | .................... |
| B | 5830 | hCoV-19/Sweden/20-07651/2020\|EPI_ISL_445236\|2020-0 | .................... | .................... | ..................... | .................... | .................... | .................... |
| B | 5831 | hCoV-19/Sweden/20-07653/2020\|EPI_ISL_445237\|2020-0 | .................... | .................... | ..................... | .................... | .................... | .................... |
| B | 5832 | hCoV-19/Sweden/20-07691/2020\|EPI_ISL_445240\|2020-0 | .................... | .................... | ..................... | .................... | .................... | .................... |
| B | 5833 | hCoV-19/Sweden/20-07741/2020\|EPI_ISL_445241\|2020-0 | .................... | .................... | ..................... | .................... | .................... | .................... |
| B | 5834 | hCoV-19/Sweden/20-07772/2020\|EPI_ISL_445242\|2020-0 | .................... | .................... | ..................... | .................... | .................... | .................... |
| B | 5835 | hCoV-19/Romania/279067/2020\|EPI_ISL_445243\|2020-04 | .................... | .................... | ..................... | .................... | .................... | .................... |
| B | 5836 | hCoV-19/Bangladesh/Akbiomed-01/2020\|EPI_ISL_445244 | .................... | .................... | ..................... | .................... | .................... | .................... |
| B | 5837 | hCoV-19/Chile/RM-ISPCH-3/2020\|EPI_ISL_445245\|2020- | .................... | .................... | ..................... | .................... | .................... | .................... |
| B | 5838 | hCoV-19/Chile/RM-ISPCH-4/2020\|EPI_ISL_445247\|2020- | .................... | .................... | ..................... | .................... | .................... | .................... |
| B | 5839 | hCoV-19/Chile/RM-ISPCH-5/2020\|EPI_ISL_445248\|2020- | .................... | .................... | ..................... | .................... | .................... | .................... |
| B | 5840 | hCoV-19/Chile/RM-ISPCH-6/2020\|EPI_ISL_445249\|2020- | .................... | .................... | ..................... | .................... | .................... | .................... |
| B | 5841 | hCoV-19/Chile/RM-ISPCH-7/2020\|EPI_ISL_445250\|2020- | .................... | .................... | ..................... | .................... | .................... | .................... |
| B | 5842 | hCoV-19/Chile/RM-ISPCH-8/2020\|EPI_ISL_445251\|2020- | .................... | .................... | ..................... | .................... | .................... | .................... |
| B | 5843 | hCoV-19/Chile/RM-ISPCH-11/2020\|EPI_ISL_445254\|2020 | .................... | .................... | ..................... | .................... | .................... | .................... |
| B | 5844 | hCoV-19/Chile/RM-ISPCH-12/2020\|EPI_ISL_445255\|2020 | .................... | .................... | ..................... | .................... | .................... | .................... |
| B | 5845 | hCoV-19/Chile/RM-ISPCH-15/2020\|EPI_ISL_445258\|2020 | .................... | .................... | ..................... | .................... | .................... | .................... |
| B | 5846 | hCoV-19/Chile/RM-ISPCH-16/2020\|EPI_ISL_445259\|2020 | .................... | .................... | ..................... | .................... | .................... | .................... |
| B | 5847 | hCoV-19/Chile/RM-ISPCH-17/2020\|EPI_ISL_445260\|2020 | .................... | .................... | ..................... | .................... | .................... | .................... |
| B | 5848 | hCoV-19/Chile/RM-ISPCH-18/2020\|EPI_ISL_445261\|2020 | .................... | .................... | ..................... | .................... | .................... | .................... |
| B | 5849 | hCoV-19/Chile/RM-ISPCH-19/2020\|EPI_ISL_445262\|2020 | .................... | .................... | ..................... | .................... | .................... | .................... |
| B | 5850 | hCoV-19/Chile/RM-ISPCH-20/2020\|EPI_ISL_445263\|2020 | .................... | .................... | ..................... | .................... | .................... | .................... |
| B | 5851 | hCoV-19/Chile/RM-ISPCH-21/2020\|EPI_ISL_445264\|2020 | .................... | .................... | ..................... | .................... | .................... | .................... |
| B | 5852 | hCoV-19/Chile/AN-ISPCH-1/2020\|EPI_ISL_445266\|2020- | .................... | .................... | ..................... | .................... | .................... | .................... |
| B | 5853 | hCoV-19/Chile/AN-ISPCH-2/2020\|EPI_ISL_445267\|2020- | .................... | .................... | ..................... | .................... | .................... | .................... |
| B | 5854 | hCoV-19/Chile/MA-ISPCH-1/2020\|EPI_ISL_445268\|2020- | .................... | .................... | ..................... | .................... | .................... | .................... |
| B | 5855 | hCoV-19/Chile/MA-ISPCH-2/2020\|EPI_ISL_445269\|2020- | .................... | .................... | ..................... | .................... | .................... | .................... |
| B | 5856 | hCoV-19/Chile/AR-ISPCH-1/2020\|EPI_ISL_445270\|2020- | .................... | .................... | ..................... | .................... | .................... | .................... |
| B | 5857 | hCoV-19/Chile/BI-ISPCH-1/2020\|EPI_ISL_445271\|2020- | .................... | .................... | ..................... | .................... | .................... | .................... |
| B | 5858 | hCoV-19/Chile/VS-ISPCH-1/2020\|EPI_ISL_445272\|2020- | .................... | .................... | ..................... | .................... | .................... | .................... |
| B | 5859 | hCoV-19/Chile/LI-ISPCH-1/2020\|EPI_ISL_445273\|2020- | .................... | .................... | ..................... | .................... | .................... | .................... |
| B | 5860 | hCoV-19/Chile/LI-ISPCH-2/2020\|EPI_ISL_445274\|2020- | .................... | .................... | ..................... | .................... | .................... | .................... |
| B | 5861 | hCoV-19/Chile/LI-ISPCH-3/2020\|EPI_ISL_445275\|2020- | .................... | .................... | ..................... | .................... | .................... | .................... |
| B | 5862 | hCoV-19/Chile/LI-ISPCH-5/2020\|EPI_ISL_445277\|2020- | .................... | .................... | ..................... | .................... | .................... | .................... |
| B | 5863 | hCoV-19/Chile/LI-ISPCH-6/2020\|EPI_ISL_445278\|2020- | .................... | .................... | ..................... | .................... | .................... | .................... |
| B | 5864 | hCoV-19/Chile/AR-ISPCH-2/2020\|EPI_ISL_445279\|2020- | .................... | .................... | ..................... | .................... | .................... | .................... |
| B | 5865 | hCoV-19/Chile/MA-ISPCH-3/2020\|EPI_ISL_445280\|2020- | .................... | .................... | ..................... | .................... | .................... | .................... |
| B | 5866 | hCoV-19/Chile/BI-ISPCH-1-2/2020\|EPI_ISL_445281\|202 | .................... | .................... | ..................... | .................... | .................... | .................... |
| B | 5867 | hCoV-19/Chile/MA-ISPCH-4/2020\|EPI_ISL_445282\|2020- | .................... | .................... | ..................... | .................... | .................... | .................... |
| B | 5868 | hCoV-19/Chile/MA-ISPCH-5/2020\|EPI_ISL_445283\|2020- | .................... | .................... | ..................... | .................... | .................... | .................... |
| B | 5869 | hCoV-19/Chile/MA-ISPCH-6/2020\|EPI_ISL_445284\|2020- | .................... | .................... | ..................... | .................... | .................... | .................... |
| B | 5870 | hCoV-19/Chile/VS-ISPCH-2/2020\|EPI_ISL_445285\|2020- | .................... | .................... | ..................... | .................... | .................... | .................... |
| B | 5871 | hCoV-19/Chile/VS-ISPCH-3/2020\|EPI_ISL_445286\|2020- | .................... | .................... | ..................... | .................... | .................... | .................... |
| B | 5872 | hCoV-19/Chile/VS-ISPCH-4/2020\|EPI_ISL_445287\|2020- | .................... | .................... | ..................... | .................... | .................... | .................... |
| B | 5873 | hCoV-19/Chile/MA-ISPCH-7/2020\|EPI_ISL_445288\|2020- | .................... | .................... | ..................... | .................... | .................... | .................... |
| B | 5874 | hCoV-19/Chile/MA-ISPCH-8/2020\|EPI_ISL_445289\|2020- | .................... | .................... | ..................... | .................... | .................... | .................... |
| B | 5875 | hCoV-19/Chile/MA-ISPCH-9/2020\|EPI_ISL_445290\|2020- | .................... | .................... | ..................... | .................... | .................... | .................... |
| B | 5876 | hCoV-19/Chile/MA-ISPCH-11/2020\|EPI_ISL_445292\|2020 | .................... | .................... | ..................... | .................... | .................... | .................... |
| B | 5877 | hCoV-19/Chile/MA-ISPCH-12/2020\|EPI_ISL_445293\|2020 | .................... | .................... | ..................... | .................... | .................... | .................... |
| B | 5878 | hCoV-19/Chile/MA-ISPCH-13/2020\|EPI_ISL_445294\|2020 | .................... | .................... | ..................... | .................... | .................... | .................... |
| B | 5879 | hCoV-19/Chile/LI-ISPCH-7/2020\|EPI_ISL_445297\|2020- | .................... | .................... | ..................... | .................... | .................... | .................... |
| B | 5880 | hCoV-19/Wales/PHWC-319ED/2020\|EPI_ISL_446514\|2020- | .................... | .................... | ..................... | .................... | .................... | .................... |
| B | 5881 | hCoV-19/Wales/PHWC-31A08/2020\|EPI_ISL_446516\|2020- | .................... | .................... | ..................... | .................... | .................... | .................... |
| B | 5882 | hCoV-19/Wales/PHWC-31A80/2020\|EPI_ISL_446522\|2020- | .................... | .................... | ..................... | .................... | .................... | .................... |
| B | 5883 | hCoV-19/Wales/PHWC-31AEA/2020\|EPI_ISL_446528\|2020- | .................... | .................... | ..................... | .................... | .................... | .................... |
| B | 5884 | hCoV-19/Wales/PHWC-31B7E/2020\|EPI_ISL_446535\|2020- | .................... | .................... | ..................... | .................... | .................... | .................... |
| B | 5885 | hCoV-19/Wales/PHWC-31EB1/2020\|EPI_ISL_446580\|2020- | .................... | .................... | ..................... | .................... | .................... | .................... |
| B | 5886 | hCoV-19/Wales/PHWC-31F54/2020\|EPI_ISL_446589\|2020- | .................... | .................... | ..................... | .................... | .................... | .................... |
| B | 5887 | hCoV-19/Wales/PHWC-31F63/2020\|EPI_ISL_446590\|2020- | .................... | .................... | ..................... | .................... | .................... | .................... |
| B | 5888 | hCoV-19/Wales/PHWC-31FBE/2020\|EPI_ISL_446594\|2020- | .................... | .................... | ..................... | .................... | .................... | .................... |
| B | 5889 | hCoV-19/Beijing/BJ390/2020\|EPI_ISL_430730\|2020-01- | .................... | .................... | ..................... | .................... | .................... | .................... |
| B | 5890 | hCoV-19/Beijing/BJ561/2020\|EPI_ISL_430731\|2020-02- | .................... | .................... | ..................... | .................... | .................... | .................... |
| B | 5891 | hCoV-19/Beijing/BJ556/2020\|EPI_ISL_430733\|2020-02- | .................... | .................... | ..................... | .................... | .................... | .................... |
| B | 5892 | hCoV-19/Beijing/BJ53/2020\|EPI_ISL_430734\|2020-01-2 | .................... | .................... | ..................... | .................... | .................... | .................... |
| B | 5893 | hCoV-19/Beijing/BJ54/2020\|EPI_ISL_430735\|2020-01-2 | .................... | .................... | ..................... | .................... | .................... | .................... |
| B | 5894 | hCoV-19/Beijing/BJ252/2020\|EPI_ISL_430736\|2020-01- | .................... | .................... | ..................... | .................... | .................... | .................... |
| B | 5895 | hCoV-19/Beijing/BJ1112/2020\|EPI_ISL_430737\|2020-02 | .................... | .................... | ..................... | .................... | .................... | .................... |
| B | 5896 | hCoV-19/Beijing/BJ589/2020\|EPI_ISL_430738\|2020-02- | .................... | .................... | ..................... | .................... | .................... | .................... |
| B | 5897 | hCoV-19/Beijing/BJ710/2020\|EPI_ISL_430740\|2020-02- | .................... | .................... | ..................... | .................... | .................... | .................... |
| B | 5898 | hCoV-19/Beijing/BJ782/2020\|EPI_ISL_430741\|2020-02- | .................... | .................... | ..................... | .................... | .................... | .................... |

| | | | | | | | | |
|---|---|---|---|---|---|---|---|---|
| B | 5899 hCoV-19/Beijing/BJ251/2020\|EPI_ISL_430742\|2020-01- | ···················· | ···················· | ···················· | ···················· | ···················· | ···················· | |
| B | 5900 hCoV-19/env/Beijing/BJ2452/2020\|EPI_ISL_430743\|202 | ···················· | ···················· | ···················· | ···················· | ···················· | ···················· | |
| B | 5901 hCoV-19/env/Beijing/BJ2454/2020\|EPI_ISL_430744\|202 | ···················· | ···················· | ···················· | ···················· | ···················· | ···················· | |
| B | 5902 hCoV-19/env/Beijing/BJ2460/2020\|EPI_ISL_430746\|202 | ···················· | ···················· | ···················· | ···················· | ···················· | ···················· | |
| B | 5903 hCoV-19/Argentina/PAIS-A0004/2020\|EPI_ISL_430796\|2 | ···················· | ···················· | ···················· | ···················· | ···················· | ···················· | |
| B | 5904 hCoV-19/Argentina/PAIS-A0005/2020\|EPI_ISL_430797\|2 | ···················· | ···················· | ···················· | ···················· | ···················· | ···················· | |
| B | 5905 hCoV-19/Argentina/PAIS-A0006/2020\|EPI_ISL_430798\|2 | ···················· | ···················· | ···················· | ···················· | ···················· | ···················· | |
| B | 5906 hCoV-19/Argentina/PAIS-A0007/2020\|EPI_ISL_430799\|2 | ···················· | ···················· | ···················· | ···················· | ···················· | ···················· | |
| B | 5907 hCoV-19/Scotland/EDB868/2020\|EPI_ISL_433260\|2020-0 | ···················· | ···················· | ···················· | ···················· | ···················· | ···················· | |
| B | 5908 hCoV-19/Scotland/EDB877/2020\|EPI_ISL_433261\|2020-0 | ···················· | ·A·················· | ···················· | ···················· | ···················· | ···················· | |
| B | 5909 hCoV-19/Scotland/EDB880/2020\|EPI_ISL_433262\|2020-0 | ···················· | ·A·················· | ···················· | ···················· | ···················· | ···················· | |
| B | 5910 hCoV-19/Scotland/EDB884/2020\|EPI_ISL_433263\|2020-0 | ···················· | ···················· | ···················· | ···················· | ···················· | ···················· | |
| B | 5911 hCoV-19/Scotland/EDB886/2020\|EPI_ISL_433265\|2020-0 | ···················· | ···················· | ···················· | ···················· | ···················· | ···················· | |
| B | 5912 hCoV-19/USA/VA-DCLS-0095/2020\|EPI_ISL_434590\|2020- | ···················· | ···················· | ···················· | ···················· | ···················· | ···················· | |
| B | 5913 hCoV-19/USA/VA-DCLS-0098/2020\|EPI_ISL_434592\|2020- | ···················· | ···················· | ···················· | ···················· | ···················· | ···················· | |
| B | 5914 hCoV-19/USA/VA-DCLS-0099/2020\|EPI_ISL_434593\|2020- | ···················· | ···················· | ···················· | ···················· | ···················· | ···················· | |
| B | 5915 hCoV-19/USA/VA-DCLS-0101/2020\|EPI_ISL_434595\|2020- | ···················· | ···················· | ···················· | ···················· | ···················· | ···················· | |
| B | 5916 hCoV-19/USA/VA-DCLS-0104/2020\|EPI_ISL_434596\|2020- | ···················· | ···················· | ···················· | ···················· | ···················· | ···················· | |
| B | 5917 hCoV-19/USA/CA-SCCPHD-UC122/2020\|EPI_ISL_435601\|20 | ···················· | ···················· | ···················· | ···················· | ···················· | ···················· | |
| B | 5918 hCoV-19/USA/CA-SCCPHD-UC129/2020\|EPI_ISL_435608\|20 | ···················· | ···················· | ···················· | ···················· | ···················· | ···················· | |
| B | 5919 hCoV-19/USA/CA-SCCPHD-UC130/2020\|EPI_ISL_435609\|20 | ···················· | ···················· | ···················· | ···················· | ···················· | ···················· | |
| B | 5920 hCoV-19/USA/CA-SCCPHD-UC133/2020\|EPI_ISL_435612\|20 | ···················· | ···················· | ···················· | ···················· | ···················· | ···················· | |
| B | 5921 hCoV-19/USA/CA-SCCPHD-UC136/2020\|EPI_ISL_435615\|20 | ···················· | ···················· | ···················· | ···················· | ···················· | ···················· | |
| B | 5922 hCoV-19/USA/CA-SCCPHD-UC142/2020\|EPI_ISL_435621\|20 | ···················· | ···················· | ···················· | ···················· | ···················· | ···················· | |
| B | 5923 hCoV-19/USA/WI-GMF-02381/2020\|EPI_ISL_436900\|2020- | ···················· | ···················· | ···················· | ···················· | ···················· | ···················· | |
| B | 5924 hCoV-19/USA/UT-01216/2020\|EPI_ISL_436903\|2020-04-0 | ···················· | ···················· | ···················· | ···················· | ···················· | ···················· | |
| B | 5925 hCoV-19/USA/UT-01218/2020\|EPI_ISL_436905\|2020-04-0 | ···················· | ···················· | ···················· | ···················· | ···················· | ···················· | |
| B | 5926 hCoV-19/USA/UT-01222/2020\|EPI_ISL_436907\|2020-04-0 | ···················· | ···················· | ···················· | ···················· | ···················· | ···················· | |
| B | 5927 hCoV-19/USA/UT-01434/2020\|EPI_ISL_436914\|2020-04-0 | ···················· | ···················· | ···················· | ···················· | ···················· | ···················· | |
| B | 5928 hCoV-19/USA/UT-01435/2020\|EPI_ISL_436915\|2020-04-1 | ···················· | ···················· | ···················· | ···················· | ···················· | ···················· | |
| B | 5929 hCoV-19/USA/UT-01437/2020\|EPI_ISL_436917\|2020-04-1 | ···················· | ···················· | ···················· | ···················· | ···················· | ···················· | |
| B | 5930 hCoV-19/USA/UT-02221/2020\|EPI_ISL_436921\|2020-04-1 | ···················· | ···················· | ···················· | ···················· | ···················· | ···················· | |
| B | 5931 hCoV-19/USA/UT-02222/2020\|EPI_ISL_436922\|2020-04-1 | ···················· | ···················· | ···················· | ···················· | ···················· | ···················· | |
| B | 5932 hCoV-19/USA/UT-02223/2020\|EPI_ISL_436923\|2020-04-1 | ···················· | ···················· | ···················· | ···················· | ···················· | ···················· | |
| B | 5933 hCoV-19/USA/UT-02224/2020\|EPI_ISL_436924\|2020-04-1 | ···················· | ···················· | ···················· | ···················· | ···················· | ···················· | |
| B | 5934 hCoV-19/USA/UT-02227/2020\|EPI_ISL_436925\|2020-04-1 | ···················· | ···················· | ···················· | ···················· | ···················· | ···················· | |
| B | 5935 hCoV-19/USA/UT-02228/2020\|EPI_ISL_436926\|2020-04-1 | ···················· | ···················· | ···················· | ···················· | ···················· | ···················· | |
| B | 5936 hCoV-19/USA/UT-02229/2020\|EPI_ISL_436927\|2020-04-1 | ···················· | ···················· | ···················· | ···················· | ···················· | ···················· | |
| B | 5937 hCoV-19/USA/UT-02231/2020\|EPI_ISL_436928\|2020-04-1 | ···················· | ···················· | ···················· | ···················· | ···················· | ···················· | |
| B | 5938 hCoV-19/USA/UT-02233/2020\|EPI_ISL_436930\|2020-04-1 | ···················· | ···················· | ···················· | ···················· | ···················· | ···················· | |
| B | 5939 hCoV-19/USA/UT-02234/2020\|EPI_ISL_436931\|2020-04-1 | ···················· | ···················· | ···················· | ···················· | ···················· | ···················· | |
| B | 5940 hCoV-19/USA/UT-02243/2020\|EPI_ISL_436934\|2020-04-1 | ···················· | ···················· | ···················· | ···················· | ···················· | ···················· | |
| B | 5941 hCoV-19/USA/UT-02244/2020\|EPI_ISL_436935\|2020-04-1 | ···················· | ···················· | ···················· | ···················· | ···················· | ···················· | |
| B | 5942 hCoV-19/USA/UT-02246/2020\|EPI_ISL_436937\|2020-04-1 | ···················· | ···················· | ···················· | ···················· | ···················· | ···················· | |
| B | 5943 hCoV-19/USA/UT-02247/2020\|EPI_ISL_436938\|2020-04-1 | ···················· | ···················· | ···················· | ···················· | ···················· | ···················· | |
| B | 5944 hCoV-19/USA/LA-BIE-071/2020\|EPI_ISL_436939\|2020-04 | ···················· | ···················· | ···················· | ···················· | ···················· | ···················· | |
| B | 5945 hCoV-19/USA/LA-BIE-074/2020\|EPI_ISL_436942\|2020-04 | ···················· | ···················· | ···················· | ···················· | ···················· | ···················· | |
| B | 5946 hCoV-19/USA/LA-BIE-075/2020\|EPI_ISL_436943\|2020-04 | ···················· | ···················· | ···················· | ···················· | ···················· | ···················· | |
| B | 5947 hCoV-19/USA/LA-BIE-076/2020\|EPI_ISL_436944\|2020-04 | ···················· | ···················· | ···················· | ···················· | ···················· | ···················· | |
| B | 5948 hCoV-19/USA/LA-BIE-077/2020\|EPI_ISL_436945\|2020-04 | ···················· | ···················· | ···················· | ···················· | ···················· | ···················· | |
| B | 5949 hCoV-19/USA/LA-BIE-078/2020\|EPI_ISL_436946\|2020-04 | ···················· | ···················· | ···················· | ···················· | ···················· | ···················· | |
| B | 5950 hCoV-19/USA/LA-BIE-079/2020\|EPI_ISL_436947\|2020-04 | ···················· | ···················· | ···················· | ···················· | ···················· | ···················· | |
| B | 5951 hCoV-19/USA/LA-BIE-080/2020\|EPI_ISL_436948\|2020-04 | ···················· | ···················· | ···················· | ···················· | ···················· | ···················· | |
| B | 5952 hCoV-19/USA/LA-BIE-081/2020\|EPI_ISL_436949\|2020-04 | ···················· | ···················· | ···················· | ···················· | ···················· | ···················· | |
| B | 5953 hCoV-19/USA/LA-BIE-082/2020\|EPI_ISL_436950\|2020-04 | ···················· | ···················· | ···················· | ···················· | ···················· | ···················· | |
| B | 5954 hCoV-19/USA/LA-BIE-084/2020\|EPI_ISL_436952\|2020-04 | ···················· | ···················· | ···················· | ···················· | ···················· | ···················· | |
| B | 5955 hCoV-19/USA/LA-BIE-085/2020\|EPI_ISL_436953\|2020-04 | ···················· | ···················· | ···················· | ···················· | ···················· | ···················· | |
| B | 5956 hCoV-19/USA/LA-BIE-086/2020\|EPI_ISL_436954\|2020-04 | ···················· | ···················· | ···················· | ···················· | ···················· | ···················· | |
| B | 5957 hCoV-19/Scotland/CVR750/2020\|EPI_ISL_439101\|2020-0 | ···················· | ···················· | ···················· | ···················· | ···················· | ···················· | |
| B | 5958 hCoV-19/Scotland/CVR756/2020\|EPI_ISL_439103\|2020-0 | ···················· | ·A·················· | ···················· | ···················· | ···················· | ···················· | |
| B | 5959 hCoV-19/Scotland/CVR763/2020\|EPI_ISL_439105\|2020-0 | ···················· | ···················· | ···················· | ···················· | ···················· | ···················· | |
| B | 5960 hCoV-19/Scotland/CVR764/2020\|EPI_ISL_439106\|2020-0 | ···················· | ···················A | ···················· | ···················· | ···················· | ···················· | 1 |
| B | 5961 hCoV-19/Scotland/CVR886/2020\|EPI_ISL_439130\|2020-0 | ···················· | ···················· | ···················· | ···················· | ···················· | ···················· | |
| B | 5962 hCoV-19/Scotland/CVR898/2020\|EPI_ISL_439132\|2020-0 | ···················· | ···················· | ···················· | ···················· | ···················· | ···················· | |
| B | 5963 hCoV-19/Scotland/CVR910/2020\|EPI_ISL_439134\|2020-0 | ···················· | ···················· | ···················· | ···················· | ···················· | ···················· | |
| B | 5964 hCoV-19/Scotland/CVR914/2020\|EPI_ISL_439135\|2020-0 | ···················· | ···················· | ···················· | ···················· | ···················· | ···················· | |
| B | 5965 hCoV-19/Scotland/EDB1059/2020\|EPI_ISL_439145\|2020- | ···················· | ·A·················· | ···················· | ···················· | ···················· | ···················· | |
| B | 5966 hCoV-19/Scotland/EDB1080/2020\|EPI_ISL_439146\|2020- | ···················· | ···················· | ···················· | ···················· | ··············N····· | ···N················ | |
| B | 5967 hCoV-19/Scotland/EDB1247/2020\|EPI_ISL_439147\|2020- | ···················· | ·A·················· | ···················· | ···················· | ···················· | ···················· | |
| B | 5968 hCoV-19/Scotland/EDB137/2020\|EPI_ISL_439148\|2020-0 | ···················· | ···················· | ···················· | ···················· | ···················· | ···················· | |
| B | 5969 hCoV-19/Scotland/EDB1992/2020\|EPI_ISL_439150\|2020- | ···················· | ·A·················· | ···················· | ···················· | ···················· | ···················· | |
| B | 5970 hCoV-19/Scotland/EDB2100/2020\|EPI_ISL_439151\|2020- | ···················· | ···················· | ···················· | ···················· | ···················· | ···················· | |
| B | 5971 hCoV-19/Scotland/EDB2101/2020\|EPI_ISL_439152\|2020- | ···················· | ···················· | ···················· | ···················· | ···················· | ···················· | |
| B | 5972 hCoV-19/Scotland/EDB2104/2020\|EPI_ISL_439153\|2020- | ···················· | ·A·················· | ···················· | ···················· | ···················· | ···················· | |
| B | 5973 hCoV-19/Scotland/EDB2105/2020\|EPI_ISL_439154\|2020- | ···················· | ···················· | ···················· | ···················· | ···················· | ···················· | |
| B | 5974 hCoV-19/Scotland/EDB2106/2020\|EPI_ISL_439155\|2020- | ···················· | ···················· | ···················· | ···················· | ···················· | ···················· | |
| B | 5975 hCoV-19/Scotland/EDB2108/2020\|EPI_ISL_439157\|2020- | ···················· | ···················· | ···················· | ···················· | ···················· | ···················· | |
| B | 5976 hCoV-19/Scotland/EDB2114/2020\|EPI_ISL_439159\|2020- | ···················· | ···················· | ···················· | ···················· | ···················· | ···················· | |

B 5977 hCoV-19/Scotland/EDB2125/2020|EPI_ISL_439160|2020- ···················· ···················· ···················· ···················· ···················· ····················
B 5978 hCoV-19/Scotland/EDB2128/2020|EPI_ISL_439161|2020- ···················· ·A·················· ···················· ···················· ···················· ····················
B 5979 hCoV-19/Scotland/EDB2130/2020|EPI_ISL_439162|2020- ···················· ·A·················· ···················· ···················· ···················· ····················
B 5980 hCoV-19/Scotland/EDB2132/2020|EPI_ISL_439164|2020- ···················· ···················· ···················· ···················· ···················· ····················
B 5981 hCoV-19/Scotland/EDB2136/2020|EPI_ISL_439166|2020- ···················· ···················· ···················· ···················· ···················· ····················
B 5983 hCoV-19/Scotland/EDB2141/2020|EPI_ISL_439168|2020- ···················· ···················· ···················· ···················· ···················· ····················
B 5984 hCoV-19/Scotland/EDB2144/2020|EPI_ISL_439170|2020- ···················· ···················· ···················· ···················· ···················· ····················
B 5985 hCoV-19/Scotland/EDB2168/2020|EPI_ISL_439171|2020- ···················· ···················· ···················· ···················· ···················· ····················
B 5986 hCoV-19/Scotland/EDB2176/2020|EPI_ISL_439172|2020- ···················· ···················· ···················· ···················· ···················· ····················
B 5987 hCoV-19/Scotland/EDB2234/2020|EPI_ISL_439173|2020- ···················· ·A·················· ···················· ···················· ···················· ····················
B 5988 hCoV-19/Scotland/EDB2242/2020|EPI_ISL_439174|2020- ···················· ·A·················· ···················· ···················· ···················· ····················
B 5989 hCoV-19/Scotland/EDB2243/2020|EPI_ISL_439175|2020- ···················· ··············C····· ···················· ···················· ···················· ····················
B 5990 hCoV-19/Scotland/EDB2244/2020|EPI_ISL_439176|2020- ···················· ···················· ···················· ···················· ···················· ····················
B 5992 hCoV-19/Scotland/EDB2268/2020|EPI_ISL_439178|2020- ···················· ·A·················· ···················· ···················· ···················· ····················
B 5993 hCoV-19/Scotland/EDB2287/2020|EPI_ISL_439179|2020- ···················· ···················· ···················· ···················· ···················· ····················
B 5994 hCoV-19/Scotland/EDB2290/2020|EPI_ISL_439180|2020- ···················· ···················· ···················· ···················· ···················· ····················
B 5995 hCoV-19/Scotland/EDB2317/2020|EPI_ISL_439181|2020- ···················· ···················· ···················· ···················· ···················· ····················
B 5996 hCoV-19/Scotland/EDB2335/2020|EPI_ISL_439182|2020- ···················· ···················· ···················· ···················· ···················· ····················
B 5997 hCoV-19/Scotland/EDB2337/2020|EPI_ISL_439183|2020- ···················· ·A·················· ···················· ···················· ···················· ····················
B 5998 hCoV-19/Scotland/EDB2339/2020|EPI_ISL_439184|2020- ···················· ···················· ···················· ···················· ···················· ····················
B 5999 hCoV-19/Scotland/EDB2341/2020|EPI_ISL_439185|2020- ···················· ···················· ···················· ···················· ···················· ····················
B 6000 hCoV-19/Scotland/EDB2357/2020|EPI_ISL_439187|2020- ···················· ···················· ···················· ···················· ···················· ····················
B 6001 hCoV-19/Scotland/EDB2358/2020|EPI_ISL_439188|2020- ···················· ···················· ···················· ···················· ···················· ····················
B 6002 hCoV-19/Scotland/EDB2359/2020|EPI_ISL_439189|2020- ···················· ·A·················· ···················· ···················· ···················· ····················
B 6003 hCoV-19/Scotland/EDB2361/2020|EPI_ISL_439190|2020- ···················· ···················· ···················· ···················· ···················· ····················
B 6004 hCoV-19/Scotland/EDB2362/2020|EPI_ISL_439191|2020- ···················· ···················· ···················· ···················· ···················· ····················
B 6005 hCoV-19/Scotland/EDB2372/2020|EPI_ISL_439193|2020- ···················· ···················· ···················· ···················· ···················· ····················
B 6006 hCoV-19/Scotland/EDB2373/2020|EPI_ISL_439194|2020- ···················· ···················· ···················· ···················· ···················· ····················
B 6007 hCoV-19/Scotland/EDB2377/2020|EPI_ISL_439195|2020- ···················· ···················· ···················· ···················· ···················· ····················
B 6008 hCoV-19/Scotland/EDB2382/2020|EPI_ISL_439196|2020- ···················· ···················· ···················· ···················· ···················· ····················
B 6009 hCoV-19/Scotland/EDB2385/2020|EPI_ISL_439197|2020- ···················· ·A·················· ···················· ···················· ···················· ····················
B 6010 hCoV-19/Scotland/EDB2389/2020|EPI_ISL_439198|2020- ···················· ·A·················· ···················· ···················· ···················· ····················
B 6011 hCoV-19/Scotland/EDB2398/2020|EPI_ISL_439199|2020- ···················· ···················· ···················· ···················· ···················· ····················
B 6012 hCoV-19/England/CAMB-72DB7/2020|EPI_ISL_440112|202 ···················· ···················· ···················· ···················· ···················· ····················
B 6013 hCoV-19/England/BRIS-121D6B/2020|EPI_ISL_440114|20 ···················· ···················· ···················· ···················· ···················· ····················
B 6014 hCoV-19/England/BRIS-121D7A/2020|EPI_ISL_440115|20 ···················· ···················· ···················· ···················· ···················· ····················
B 6015 hCoV-19/England/BRIS-1222B6/2020|EPI_ISL_440116|20 ···················· ···················· ···················· ···················· ···················· ····················
B 6016 hCoV-19/England/CAMB-733B4/2020|EPI_ISL_440117|202 ···················· ···················· ···················· ···················· ···················· ····················
B 6017 hCoV-19/England/CAMB-72D20/2020|EPI_ISL_440120|202 ···················· ···················· ···················· ···················· ···················· ····················
B 6018 hCoV-19/England/CAMB-72D6C/2020|EPI_ISL_440121|202 ···················· ···················· ···················· ···················· ···················· ····················
B 6019 hCoV-19/England/CAMB-72E3C/2020|EPI_ISL_440122|202 ···················· ···················· ···················· ···················· ···················· ····················
B 6020 hCoV-19/England/BRIS-121D01/2020|EPI_ISL_440123|20 ···················· ···················· ···················· ···················· ···················· ····················
B 6021 hCoV-19/England/BRIS-122034/2020|EPI_ISL_440124|20 ···················· ···················· ···················· ···················· ···················· ····················
B 6022 hCoV-19/England/CAMB-7324E/2020|EPI_ISL_440132|202 ···················· ···················· ···················· ···················· ···················· ····················
B 6023 hCoV-19/England/BRIS-1217B8/2020|EPI_ISL_440135|20 ···················· ···················· ···················· ···················· ···················· ····················
B 6024 hCoV-19/England/CAMB-73220/2020|EPI_ISL_440138|202 ···················· ···················· ···················· ···················· ···················· ····················
B 6025 hCoV-19/England/BRIS-121C31/2020|EPI_ISL_440139|20 ···················· ···················· ···················· ···················· ···················· ····················
B 6026 hCoV-19/England/BRIS-1219FE/2020|EPI_ISL_440141|20 ···················· ···················· ···················· ···················· ···················· ····················
B 6027 hCoV-19/England/CAMB-731D8/2020|EPI_ISL_440143|202 ···················· ···················· ···················· ···················· ···················· ····················
B 6028 hCoV-19/England/CAMB-72FEE/2020|EPI_ISL_440144|202 ···················· ···················· ···················· ···················· ···················· ····················
B 6029 hCoV-19/England/CAMB-730BD/2020|EPI_ISL_440146|202 ···················· ···················· ···················· ···················· ···················· ····················
B 6030 hCoV-19/England/CAMB-73581/2020|EPI_ISL_440147|202 ···················· ···················· ···················· ···················· ···················· ····················
B 6031 hCoV-19/England/BRIS-121B8F/2020|EPI_ISL_440149|20 ···················· ···················· ···················· ···················· ···················· ····················
B 6032 hCoV-19/England/CAMB-72FB1/2020|EPI_ISL_440150|202 ···················· ···················· ···················· ···················· ···················· ····················
B 6033 hCoV-19/England/CAMB-73299/2020|EPI_ISL_440152|202 ···················· ···················· ···················· ···················· ···················· ····················
B 6034 hCoV-19/England/CAMB-73615/2020|EPI_ISL_440155|202 ···················· ···················· ···················· ···················· ···················· ····················
B 6035 hCoV-19/England/BRIS-1218D3/2020|EPI_ISL_440156|20 ···················· ···················· ···················· ···················· ···················· ····················
B 6036 hCoV-19/England/BRIS-121A91/2020|EPI_ISL_440158|20 ···················· ···················· ···················· ···················· ···················· ····················
B 6037 hCoV-19/England/CAMB-736AC/2020|EPI_ISL_440159|202 ···················· ···················· ···················· ···················· ···················· ····················
B 6038 hCoV-19/England/CAMB-72FA2/2020|EPI_ISL_440161|202 ···················· ···················· ···················· ···················· ···················· ····················
B 6039 hCoV-19/England/BRIS-1218C4/2020|EPI_ISL_440162|20 ···················· ·A·················· ···················· ···················· ···················· ····················
B 6040 hCoV-19/England/CAMB-73712/2020|EPI_ISL_440163|202 ···················· ···················· ···················· ···················· ···················· ····················
B 6041 hCoV-19/England/CAMB-735AF/2020|EPI_ISL_440164|202 ···················· ···················· ···················· ···················· ···················· ····················
B 6042 hCoV-19/England/CAMB-73053/2020|EPI_ISL_440166|202 ···················· ···················· ···················· ···················· ···················· ····················
B 6043 hCoV-19/England/BRIS-121BE9/2020|EPI_ISL_440167|20 ···················· ···················· ···················· ···················· ···················· ····················
B 6044 hCoV-19/England/BRIS-121AA0/2020|EPI_ISL_440168|20 ···················· ···················· ···················· ···················· ···················· ····················
B 6045 hCoV-19/England/CAMB-73026/2020|EPI_ISL_440169|202 ···················· ···················· ···················· ···················· ···················· ····················
B 6046 hCoV-19/England/CAMB-73202/2020|EPI_ISL_440170|202 ···················· ···················· ···················· ···················· ···················· ····················
B 6047 hCoV-19/England/CAMB-7325d/2020|EPI_ISL_440176|202 ···················· ···················· ···················· ···················· ···················· ····················
B 6048 hCoV-19/England/CAMB-73150/2020|EPI_ISL_440177|202 ···················· ···················· ···················· ···················· ···················· ····················
B 6049 hCoV-19/England/BRIS-121879/2020|EPI_ISL_440178|20 ···················· ···················· ···················· ···················· ···················· ····················
B 6050 hCoV-19/England/CAMB-730AE/2020|EPI_ISL_440179|202 ···················· ···················· ···················· ···················· ···················· ····················
B 6051 hCoV-19/England/CAMB-7341B/2020|EPI_ISL_440181|202 ···················· ···················· ···················· ···················· ···················· ····················
B 6052 hCoV-19/England/CAMB-72F75/2020|EPI_ISL_440182|202 ···················· ···················· ···················· ···················· ···················· ····················
B 6053 hCoV-19/England/CAMB-73062/2020|EPI_ISL_440183|202 ···················· ···················· ···················· ···················· ···················· ····················
B 6054 hCoV-19/England/CAMB-7327B/2020|EPI_ISL_440184|202 ···················· ···················· ···················· ···················· ···················· ····················
B 6055 hCoV-19/England/CAMB-7369D/2020|EPI_ISL_440185|202 ···················· ···················· ···················· ···················· ···················· ····················
B 6056 hCoV-19/England/CAMB-73518/2020|EPI_ISL_440186|202 ···················· ···················· ···················· ···················· ···················· ····················

| | | | | | | | |
|---|---|---|---|---|---|---|---|
| B | 6057 hCoV-19/England/BRIS-12185B/2020\|EPI_ISL_440187\|20 | ···················· | ···················· | ···················· | ···················· | ···················· | ···················· |
| B | 6058 hCoV-19/England/CAMB-72F57/2020\|EPI_ISL_440188\|202 | ···················· | ···················· | ···················· | ···················· | ···················· | ···················· |
| B | 6059 hCoV-19/England/BRIS-1217E5/2020\|EPI_ISL_440189\|20 | ···················· | ···················· | ···················· | ···················· | ···················· | ···················· |
| B | 6060 hCoV-19/England/BRIS-121B34/2020\|EPI_ISL_440194\|20 | ···················· | ···················· | ···················· | ···················· | ···················· | ···················· |
| B | 6061 hCoV-19/England/CAMB-731E7/2020\|EPI_ISL_440195\|202 | ···················· | ···················· | ···················· | ···················· | ···················· | ···················· |
| B | 6062 hCoV-19/England/BRIS-121994/2020\|EPI_ISL_440196\|20 | ···················· | ·············A······ | ···················· | ···················· | ···················· | ···················· |
| B | 6063 hCoV-19/England/BRIS-121BAD/2020\|EPI_ISL_440198\|20 | ···················· | ···················· | ···················· | ···················· | ···················· | ···················· |
| B | 6066 hCoV-19/Northern Ireland/NIRE-102322/2020\|EPI_ISL_ | ···················· | ···················· | ···················· | ···················· | ···················· | ···················· |
| B | 6067 hCoV-19/Northern Ireland/NIRE-10235F/2020\|EPI_ISL_ | ···················· | ···················· | ···················· | ···················· | ···················· | ···················· |
| B | 6068 hCoV-19/Northern Ireland/NIRE-10236E/2020\|EPI_ISL_ | ···················· | ···················· | ···················· | ···················· | ··N················· | ···················· |
| B | 6069 hCoV-19/Northern Ireland/NIRE-10237D/2020\|EPI_ISL_ | ···················· | ···················· | ···················· | ···················· | ···················· | ···················· |
| B | 6071 hCoV-19/Northern Ireland/NIRE-1023AA/2020\|EPI_ISL_ | ···················· | ···················· | ···················· | ···················· | ···················· | ···················· |
| B | 6072 hCoV-19/Northern Ireland/NIRE-1023C8/2020\|EPI_ISL_ | ···················· | ···················· | ···················· | ···················· | ···················· | ···················· |
| B | 6073 hCoV-19/Northern Ireland/NIRE-1023D7/2020\|EPI_ISL_ | ···················· | ···················· | ···················· | ···················· | ···················· | ···················· |
| B | 6074 hCoV-19/Northern Ireland/NIRE-1023E6/2020\|EPI_ISL_ | ···················· | ···················· | ···················· | ···················· | ···················· | ···················· |
| B | 6075 hCoV-19/Northern Ireland/NIRE-1023F5/2020\|EPI_ISL_ | ···················· | ···················· | ···················· | ···················· | ···················· | ···················· |
| B | 6076 hCoV-19/Northern Ireland/NIRE-FB378/2020\|EPI_ISL_4 | ···················· | ···················· | ···················· | ···················· | ···················· | ···················· |
| B | 6077 hCoV-19/Northern Ireland/NIRE-FB396/2020\|EPI_ISL_4 | ···················· | ···················· | ···················· | ···················· | ···················· | ···················· |
| B | 6078 hCoV-19/Northern Ireland/NIRE-FB3B4/2020\|EPI_ISL_4 | ···················· | ···················· | ···················· | ···················· | ···················· | ···················· |
| B | 6079 hCoV-19/Northern Ireland/NIRE-FB3D2/2020\|EPI_ISL_4 | ···················· | ···················· | ···················· | ···················· | ···················· | ···················· |
| B | 6080 hCoV-19/Northern Ireland/NIRE-FB3E1/2020\|EPI_ISL_4 | ···················· | ···················· | ···················· | ···················· | ···················· | ···················· |
| B | 6081 hCoV-19/Northern Ireland/NIRE-FB3F0/2020\|EPI_ISL_4 | ···················· | ···················· | ···················· | ···················· | ···················· | ···················· |
| B | 6082 hCoV-19/Northern Ireland/NIRE-FB40C/2020\|EPI_ISL_4 | ···················· | ···················· | ···················· | ···················· | ···················· | ···················· |
| B | 6083 hCoV-19/England/NOTT-110084/2020\|EPI_ISL_441465\|20 | ···················· | ···················· | ···················· | ···················· | ···················· | ···················· |
| B | 6084 hCoV-19/England/NOTT-110163/2020\|EPI_ISL_441478\|20 | ···················· | ···················· | ···················· | ···················· | ···················· | ···················· |
| B | 6085 hCoV-19/England/NOTT-110172/2020\|EPI_ISL_441479\|20 | ···················· | ···················· | ···················· | ···················· | ···················· | ···················· |
| B | 6086 hCoV-19/England/BRIS-1234BF/2020\|EPI_ISL_442710\|20 | ···················· | ···················· | ···················· | ···················· | ···················· | ···················· |
| B | 6087 hCoV-19/England/BRIS-1233D0/2020\|EPI_ISL_442711\|20 | ···················· | ···················· | ···················· | ···················· | ···················· | ···················· |
| B | 6088 hCoV-19/England/BRIS-1239ED/2020\|EPI_ISL_442712\|20 | ···················· | ···················· | ···················· | ···················· | ···················· | ···················· |
| B | 6089 hCoV-19/England/BRIS-123543/2020\|EPI_ISL_442713\|20 | ···················· | ···················· | ···················· | ···················· | ···················· | ···················· |
| B | 6090 hCoV-19/England/BRIS-123BBA/2020\|EPI_ISL_442714\|20 | ···················· | ····A··············· | ···················· | ···················· | ···················· | ···················· |
| B | 6091 hCoV-19/England/BRIS-1238B3/2020\|EPI_ISL_442718\|20 | ···················· | ····A··············· | ···················· | ···················· | ···················· | ···················· |
| B | 6092 hCoV-19/England/BRIS-123103/2020\|EPI_ISL_442719\|20 | ···················· | ···················· | ···················· | ···················· | ···················· | ···················· |
| B | 6093 hCoV-19/England/BRIS-123482/2020\|EPI_ISL_442720\|20 | ···················· | ···················· | ···················· | ···················· | ···················· | ···················· |
| B | 6094 hCoV-19/England/BRIS-123AEA/2020\|EPI_ISL_442722\|20 | ···················· | ···················· | ···················· | ···················· | ···················· | ···················· |
| B | 6095 hCoV-19/England/BRIS-123130/2020\|EPI_ISL_442724\|20 | ···················· | ···················· | ···················· | ···················· | ···················· | ···················· |
| B | 6096 hCoV-19/England/BRIS-123279/2020\|EPI_ISL_442725\|20 | ···················· | ···················· | ···················· | ···················· | ···················· | ···················· |
| B | 6097 hCoV-19/England/BRIS-1235AD/2020\|EPI_ISL_442726\|20 | ···················· | ···················· | ···················· | ···················· | ···················· | ···················· |
| B | 6098 hCoV-19/England/BRIS-1234A0/2020\|EPI_ISL_442727\|20 | ···················· | ···················· | ···················· | ···················· | ···················· | ···················· |
| B | 6099 hCoV-19/England/BRIS-1230AC/2020\|EPI_ISL_442728\|20 | ···················· | ···················· | ···················· | ···················· | ···················· | ···················· |
| B | 6100 hCoV-19/England/BRIS-1239FC/2020\|EPI_ISL_442729\|20 | ···················· | ···················· | ···················· | ···················· | ···················· | ···················· |
| B | 6101 hCoV-19/England/BRIS-123112/2020\|EPI_ISL_442730\|20 | ···················· | ···················· | ···················· | ···················· | ···················· | ···················· |
| B | 6102 hCoV-19/England/BRIS-1236E6/2020\|EPI_ISL_442731\|20 | ···················· | ···················· | ···················· | ···················· | ···················· | ···················· |
| B | 6103 hCoV-19/England/BRIS-1234EC/2020\|EPI_ISL_442732\|20 | ···················· | ···················· | ···················· | ···················· | ···················· | ···················· |
| B | 6104 hCoV-19/England/BRIS-12359E/2020\|EPI_ISL_442734\|20 | ···················· | ···················· | ···················· | ···················· | ···················· | ···················· |
| B | 6105 hCoV-19/England/BRIS-123121/2020\|EPI_ISL_442735\|20 | ···················· | ···················· | ···················· | ···················· | ···················· | ···················· |
| B | 6106 hCoV-19/England/BRIS-123051/2020\|EPI_ISL_442736\|20 | ···················· | ···················· | ···················· | ···················· | ···················· | ···················· |
| B | 6107 hCoV-19/England/BRIS-12369B/2020\|EPI_ISL_442737\|20 | ···················· | ···················· | ···················· | ···················· | ···················· | ···················· |
| B | 6108 hCoV-19/England/BRIS-12307F/2020\|EPI_ISL_442739\|20 | ···················· | ···················· | ···················· | ···················· | ···················· | ···················· |
| B | 6109 hCoV-19/England/BRIS-1235CB/2020\|EPI_ISL_442740\|20 | ···················· | ···················· | ···················· | ···················· | ···················· | ···················· |
| B | 6110 hCoV-19/England/BRIS-12314F/2020\|EPI_ISL_442741\|20 | ···················· | ···················· | ···················· | ···················· | ···················· | ···················· |
| B | 6111 hCoV-19/England/BRIS-123024/2020\|EPI_ISL_442742\|20 | ···················· | ···················· | ···················· | ···················· | ···················· | ···················· |
| B | 6112 hCoV-19/England/BRIS-123BD8/2020\|EPI_ISL_442743\|20 | ···················· | ···················· | ···················· | ···················· | ···················· | ···················· |
| B | 6113 hCoV-19/England/BRIS-12323D/2020\|EPI_ISL_442744\|20 | ···················· | ···················· | ···················· | ···················· | ···················· | ···················· |
| B | 6114 hCoV-19/England/BRIS-123B05/2020\|EPI_ISL_442745\|20 | ···················· | ···················· | ···················· | ···················· | ···················· | ···················· |
| B | 6115 hCoV-19/England/BRIS-123974/2020\|EPI_ISL_442747\|20 | ···················· | ···················· | ···················· | ···················· | ···················· | ···················· |
| B | 6116 hCoV-19/England/BRIS-1230D9/2020\|EPI_ISL_442748\|20 | ···················· | ···················· | ···················· | ···················· | ···················· | ···················· |
| B | 6117 hCoV-19/England/BRIS-123622/2020\|EPI_ISL_442749\|20 | ···················· | ···················· | ···················· | ···················· | ···················· | ···················· |
| B | 6118 hCoV-19/England/BRIS-1238D1/2020\|EPI_ISL_442751\|20 | ···················· | ···················· | ···················· | ···················· | ···················· | ···················· |
| B | 6119 hCoV-19/England/BRIS-123A08/2020\|EPI_ISL_442774\|20 | ···················· | ···················· | ···················· | ···················· | ···················· | ···················· |
| B | 6120 hCoV-19/England/BRIS-123491/2020\|EPI_ISL_442775\|20 | ···················· | ···················· | ···················· | ···················· | ···················· | ···················· |
| B | 6121 hCoV-19/England/BRIS-1236C8/2020\|EPI_ISL_442777\|20 | ···················· | ···················· | ···················· | ···················· | ···················· | ···················· |
| B | 6122 hCoV-19/England/BRIS-1231B8/2020\|EPI_ISL_442778\|20 | ···················· | ···················· | ···················· | ···················· | ···················· | ···················· |
| B | 6123 hCoV-19/England/BRIS-123C3F/2020\|EPI_ISL_442779\|20 | ···················· | ···················· | ···················· | ···················· | ···················· | ···················· |
| B | 6124 hCoV-19/England/BRIS-123C5D/2020\|EPI_ISL_442780\|20 | ···················· | ···················· | ···················· | ···················· | ···················· | ···················· |
| B | 6125 hCoV-19/England/BRIS-123516/2020\|EPI_ISL_442781\|20 | ···················· | ···················· | ···················· | ···················· | ···················· | ···················· |
| B | 6126 hCoV-19/England/BRIS-12390B/2020\|EPI_ISL_442782\|20 | ········T··········· | ···················· | ···················· | ···················· | ···················· | ···················· |
| B | 6127 hCoV-19/England/BRIS-12322E/2020\|EPI_ISL_442783\|20 | ···················· | ···················· | ···················· | ···················· | ···················· | ···················· |
| B | 6128 hCoV-19/England/BRIS-12377A/2020\|EPI_ISL_442784\|20 | ···················· | ···················· | ···················· | ···················· | ···················· | ···················· |
| B | 6129 hCoV-19/England/BRIS-1238C2/2020\|EPI_ISL_442785\|20 | ···················· | ···················· | ···················· | ···················· | ···················· | ···················· |
| B | 6130 hCoV-19/England/BRIS-1231E5/2020\|EPI_ISL_442787\|20 | ···················· | ···················· | ···················· | ···················· | ···················· | ···················· |
| B | 6131 hCoV-19/England/BRIS-12333A/2020\|EPI_ISL_442788\|20 | ···················· | ···················· | ···················· | ···················· | ···················· | ···················· |
| B | 6132 hCoV-19/England/BRIS-123B23/2020\|EPI_ISL_442790\|20 | ···················· | ····A··············· | ···················· | ···················· | ···················· | ···················· |
| B | 6133 hCoV-19/England/BRIS-123358/2020\|EPI_ISL_442791\|20 | ···················· | ···················· | ···················· | ···················· | ···················· | ···················· |
| B | 6134 hCoV-19/England/BRIS-1233A3/2020\|EPI_ISL_442792\|20 | ···················· | ···················· | ···················· | ···················· | ···················· | ···················· |
| B | 6135 hCoV-19/England/BRIS-123956/2020\|EPI_ISL_442793\|20 | ···················· | ···················· | ···················· | ···················· | ···················· | ···················· |
| B | 6136 hCoV-19/England/BRIS-1231C7/2020\|EPI_ISL_442794\|20 | ···················· | ···················· | ···················· | ···················· | ···················· | ···················· |
| B | 6137 hCoV-19/England/BRIS-1230BB/2020\|EPI_ISL_442795\|20 | ···················· | ···················· | ···················· | ···················· | ···················· | ···················· |

| | | | | | | | | | |
|---|---|---|---|---|---|---|---|---|---|
| B | 6138 | hCoV-19/England/BRIS-123C02/2020\|EPI_ISL_442796\|20 | ···················· | ···················· | ···················· | ···················· | ···················· | ···················· | |
| B | 6139 | hCoV-19/England/BRIS-123A62/2020\|EPI_ISL_442797\|20 | ···················· | ···················· | ···················· | ···················· | ···················· | ···················· | |
| B | 6140 | hCoV-19/England/BRIS-1238E0/2020\|EPI_ISL_442799\|20 | ···················· | ···················· | ···················· | ···················· | ···················· | ···················· | |
| B | 6141 | hCoV-19/Sweden/20-07666/2020\|EPI_ISL_445238\|2020-0 | ···················· | ···················· | ···················· | ···················· | ···················· | ···················· | |
| B | 6142 | hCoV-19/Sweden/20-07672/2020\|EPI_ISL_445239\|2020-0 | ···················· | ···················· | ···················· | ···················· | ···················· | ···················· | |
| B | 6143 | hCoV-19/Greece/235_31783/2020\|EPI_ISL_529147\|2020- | ···················· | ···················· | ···················· | ···················· | ···················· | ···················· | |
| B | 6144 | hCoV-19/Greece/45_35378/2020\|EPI_ISL_529148\|2020-0 | ···················· | ···················· | ···················· | ···················· | ···················· | ···················· | |
| B | 6147 | hCoV-19/Australia/VIC1305/2020\|EPI_ISL_430705\|2020 | ···················· | ···················· | ···················· | ···················· | ···················· | ···················· | |
| B | 6148 | hCoV-19/Australia/VIC1345/2020\|EPI_ISL_430708\|2020 | ···················· | ···················· | ···················· | ···················· | ···················· | ···················· | |
| B | 6150 | hCoV-19/Australia/VIC1313/2020\|EPI_ISL_430714\|2020 | ···················· | ···················· | ···················· | ···················· | ···················· | ·R·················· | |
| B | 6152 | hCoV-19/Spain/MD-H12-16/2020\|EPI_ISL_430718\|2020-0 | ···················· | ···················· | ···················· | ···················· | ···················· | ···················· | |
| B | 6153 | hCoV-19/Spain/MD-H12-LP29/2020\|EPI_ISL_430719\|2020 | ···················· | ···················· | ···················· | --------············ | ···················· | ···················· | 1 |
| B | 6154 | hCoV-19/Spain/MD-H12-LP32/2020\|EPI_ISL_430720\|2020 | ···················· | ···················· | ···················· | ···················· | ···················· | ···················· | |
| B | 6155 | hCoV-19/Spain/MD-H12-LP33/2020\|EPI_ISL_430721\|2020 | ···················· | ···················· | ···················· | ···················· | ···················· | ···················· | |
| B | 6156 | hCoV-19/Beijing/BJ287/2020\|EPI_ISL_430722\|2020-01- | ···················· | ···················· | ···················· | ···················· | ···················· | ···················· | |
| B | 6157 | hCoV-19/Beijing/BJ752/2020\|EPI_ISL_430724\|2020-02- | ···················· | ···················· | ···················· | ···················· | ···················· | ···················· | |
| B | 6158 | hCoV-19/Beijing/BJ288/2020\|EPI_ISL_430725\|2020-01- | ···················· | ···················· | ···················· | ···················· | ···················· | ···················· | |
| B | 6159 | hCoV-19/Beijing/BJ492/2020\|EPI_ISL_430728\|2020-02- | ···················· | ···················· | ···················· | ···················· | ···················· | ···················· | |
| B | 6160 | hCoV-19/Beijing/BJ709/2020\|EPI_ISL_430729\|2020-02- | ···················· | ···················· | ···················· | ···················· | ···················· | ···················· | |
| B | 6161 | hCoV-19/USA/CA-CZB088/2020\|EPI_ISL_430791\|2020-03- | ···················· | ···················· | ···················· | ···················· | ···················· | ···················· | |
| B | 6162 | hCoV-19/USA/CA-CZB0716/2020\|EPI_ISL_430792\|2020-03 | ···················· | ···················· | ···················· | ···················· | ···················· | ···················· | |
| B | 6163 | hCoV-19/USA/WA-S440/2020\|EPI_ISL_434200\|2020-04-01 | ···················· | ···················· | ···················· | ···················· | ···················· | ···················· | |
| B | 6164 | hCoV-19/USA/WA-S441/2020\|EPI_ISL_434201\|2020-03-31 | ···················· | ···················· | ···················· | ···················· | ···················· | ···················· | |
| B | 6165 | hCoV-19/USA/WA-S442/2020\|EPI_ISL_434202\|2020-03-26 | ···················· | ···················· | ···················· | ···················· | ···················· | ···················· | |
| B | 6166 | hCoV-19/USA/WA-S444/2020\|EPI_ISL_434204\|2020-04-01 | ···················· | ···················· | ···················· | ···················· | ···················· | ···················· | |
| B | 6167 | hCoV-19/USA/WA-S445/2020\|EPI_ISL_434205\|2020-04-01 | ···················· | ···················· | ···················· | ···················· | ···················· | ···················· | |
| B | 6168 | hCoV-19/USA/WA-S446/2020\|EPI_ISL_434206\|2020-04-01 | ···················· | ···················· | ···················· | ···················· | ···················· | ···················· | |
| B | 6169 | hCoV-19/USA/WA-S447/2020\|EPI_ISL_434207\|2020-04-01 | ···················· | ···················· | ···················· | ···················· | ···················· | ···················· | |
| B | 6170 | hCoV-19/USA/WA-S448/2020\|EPI_ISL_434208\|2020-04-01 | ···················· | ···················· | ···················· | ···················· | ···················· | ···················· | |
| B | 6171 | hCoV-19/USA/WA-S449/2020\|EPI_ISL_434209\|2020-03-31 | ···················· | ···················· | ···················· | ···················· | ···················· | ···················· | |
| B | 6172 | hCoV-19/USA/WA-S451/2020\|EPI_ISL_434211\|2020-03-31 | ···················· | ···················· | ···················· | ···················· | ···················· | ···················· | |
| B | 6173 | hCoV-19/USA/WA-S452/2020\|EPI_ISL_434212\|2020-03-31 | ···················· | ···················· | ···················· | ···················· | ···················· | ···················· | |
| B | 6174 | hCoV-19/USA/WA-S453/2020\|EPI_ISL_434213\|2020-03-30 | ···················· | ···················· | ···················· | ···················· | ···················· | ···················· | |
| B | 6175 | hCoV-19/USA/WA-S454/2020\|EPI_ISL_434214\|2020-03-31 | ···················· | ···················· | ···················· | ···················· | ···················· | ···················· | |
| B | 6176 | hCoV-19/USA/WA-S455/2020\|EPI_ISL_434215\|2020-03-31 | ···················· | ···················· | ···················· | ···················· | ···················· | ···················· | |
| B | 6177 | hCoV-19/USA/WA-S478/2020\|EPI_ISL_434238\|2020-04-01 | ···················· | ···················· | ···················· | ···················· | ···················· | ···················· | |
| B | 6178 | hCoV-19/USA/WA-S479/2020\|EPI_ISL_434239\|2020-04-04 | ···················· | ···················· | ···················· | ···················· | ···················· | ···················· | |
| B | 6179 | hCoV-19/USA/WA-S481/2020\|EPI_ISL_434241\|2020-04-02 | ···················· | ···················· | ···················· | ···················· | ···················· | ···················· | |
| B | 6180 | hCoV-19/USA/WA-S482/2020\|EPI_ISL_434242\|2020-04-06 | ···················· | ···················· | ···················· | ···················· | ···················· | ···················· | |
| B | 6181 | hCoV-19/USA/WA-S483/2020\|EPI_ISL_434243\|2020-04-05 | ···················· | ···················· | ···················· | ···················· | ···················· | ···················· | |
| B | 6182 | hCoV-19/USA/WA-S484/2020\|EPI_ISL_434244\|2020-04-02 | ···················· | ···················· | ···················· | ···················· | ···················· | ···················· | |
| B | 6183 | hCoV-19/USA/WA-S485/2020\|EPI_ISL_434245\|2020-04-01 | ···················· | ···················· | ···················· | ···················· | ···················· | ···················· | |
| B | 6184 | hCoV-19/USA/WA-S486/2020\|EPI_ISL_434246\|2020-04-01 | ···················· | ···················· | ···················· | ···················· | ···················· | ···················· | |
| B | 6185 | hCoV-19/USA/WA-S487/2020\|EPI_ISL_434247\|2020-03-31 | ···················· | ···················· | ···················· | ···················· | ···················· | ···················· | |
| B | 6186 | hCoV-19/USA/WA-S488/2020\|EPI_ISL_434248\|2020-04-01 | ········T··········· | ···················· | ···················· | ···················· | ···················· | ···················· | |
| B | 6187 | hCoV-19/USA/WA-S500/2020\|EPI_ISL_434260\|2020-03-25 | ···················· | ···················· | ···················· | ···················· | ···················· | ···················· | |
| B | 6188 | hCoV-19/USA/WA-S502/2020\|EPI_ISL_434262\|2020-03-24 | ···················· | ···················· | ···················· | ···················· | ···················· | ···················· | |
| B | 6189 | hCoV-19/USA/WA-S503/2020\|EPI_ISL_434263\|2020-03-25 | ···················· | ···················· | ···················· | ···················· | ···················· | ···················· | |
| B | 6190 | hCoV-19/USA/WA-S504/2020\|EPI_ISL_434264\|2020-04-01 | ···················· | ···················· | ···················· | ···················· | ···················· | ···················· | |
| B | 6191 | hCoV-19/USA/WA-S505/2020\|EPI_ISL_434265\|2020-03-29 | ···················· | ···················· | ···················· | ···················· | ···················· | ···················· | |
| B | 6192 | hCoV-19/USA/WA-S506/2020\|EPI_ISL_434266\|2020-04-01 | ···················· | ···················· | ···················· | ···················· | ···················· | ···················· | |
| B | 6193 | hCoV-19/USA/WA-S508/2020\|EPI_ISL_434268\|2020-03-30 | ···················· | ···················· | ···················· | ···················· | ···················· | ···················· | |
| B | 6194 | hCoV-19/USA/WA-S509/2020\|EPI_ISL_434269\|2020-03-30 | ···················· | ···················· | ···················· | ···················· | ···················· | ···················· | |
| B | 6195 | hCoV-19/USA/WA-S510/2020\|EPI_ISL_434270\|2020-03-25 | ···················· | ···················· | ···················· | ···················· | ···················· | ···················· | |
| B | 6196 | hCoV-19/USA/WA-S511/2020\|EPI_ISL_434271\|2020-03-24 | ···················· | ···················· | ···················· | ···················· | ···················· | ···················· | |
| B | 6197 | hCoV-19/USA/WA-S512/2020\|EPI_ISL_434272\|2020-04-01 | ···················· | ···················· | ···················· | ···················· | ···················· | ···················· | |
| B | 6198 | hCoV-19/USA/WA-S513/2020\|EPI_ISL_434273\|2020-04-01 | ···················· | ···················· | ···················· | ···················· | ···················· | ···················· | |
| B | 6199 | hCoV-19/USA/WA-S514/2020\|EPI_ISL_434274\|2020-03-29 | ···················· | ···················· | ···················· | ···················· | ···················· | ···················· | |
| B | 6200 | hCoV-19/USA/WA-S516/2020\|EPI_ISL_434276\|2020-04-01 | ···················· | ···················· | ···················· | ···················· | ···················· | ···················· | |
| B | 6201 | hCoV-19/USA/WA-S517/2020\|EPI_ISL_434277\|2020-03-24 | ···················· | ···················· | ···················· | ···················· | ···················· | ···················· | |
| B | 6202 | hCoV-19/USA/WA-S519/2020\|EPI_ISL_434279\|2020-03-27 | ···················· | ···················· | ···················· | ···················· | ···················· | ···················· | |
| B | 6203 | hCoV-19/USA/WA-S520/2020\|EPI_ISL_434280\|2020-03-31 | ···················· | ···················· | ···················· | ···················· | ···················· | ···················· | |
| B | 6204 | hCoV-19/USA/WA-S521/2020\|EPI_ISL_434281\|2020-03-25 | ···················· | ···················· | ···················· | ···················· | ···················· | ···················· | |
| B | 6205 | hCoV-19/USA/WA-S522/2020\|EPI_ISL_434282\|2020-04-01 | ···················· | ···················· | ···················· | ···················· | ···················· | ···················· | |
| B | 6206 | hCoV-19/USA/WA-S523/2020\|EPI_ISL_434283\|2020-03-26 | ···················· | ···················· | ···················· | ···················· | ···················· | ···················· | |
| B | 6207 | hCoV-19/USA/WA-S524/2020\|EPI_ISL_434284\|2020-04-01 | ···················· | ···················· | ···················· | ···················· | ···················· | ···················· | |
| B | 6208 | hCoV-19/USA/WA-S525/2020\|EPI_ISL_434285\|2020-04-01 | ···················· | ···················· | ···················· | ···················· | ···················· | ···················· | |
| B | 6209 | hCoV-19/USA/WA-S526/2020\|EPI_ISL_434286\|2020-03-31 | ···················· | ···················· | ···················· | ···················· | ···················· | ···················· | |
| B | 6210 | hCoV-19/USA/WA-S527/2020\|EPI_ISL_434287\|2020-03-31 | ···················· | ···················· | ···················· | ···················· | ···················· | ···················· | |
| B | 6211 | hCoV-19/USA/WA-S528/2020\|EPI_ISL_434288\|2020-03-31 | ···················· | ···················· | ···················· | ···················· | ···················· | ···················· | |
| B | 6212 | hCoV-19/USA/WA-S529/2020\|EPI_ISL_434289\|2020-03-24 | ···················· | ···················· | ···················· | ···················· | ···················· | ···················· | |
| B | 6213 | hCoV-19/USA/WA-S530/2020\|EPI_ISL_434290\|2020-03-24 | ···················· | ···················· | ···················· | ···················· | ···················· | ···················· | |
| B | 6214 | hCoV-19/USA/WA-S531/2020\|EPI_ISL_434291\|2020-03-24 | ···················· | ···················· | ···················· | ···················· | ···················· | ···················· | |
| B | 6215 | hCoV-19/USA/WA-S533/2020\|EPI_ISL_434293\|2020-03-25 | ···················· | ···················· | ···················· | ···················· | ···················· | ···················· | |
| B | 6216 | hCoV-19/USA/WA-S534/2020\|EPI_ISL_434294\|2020-03-25 | ···················· | ···················· | ···················· | ···················· | ···················· | ···················· | |
| B | 6217 | hCoV-19/USA/WA-S536/2020\|EPI_ISL_434296\|2020-03-24 | ···················· | ···················· | ···················· | ···················· | ···················· | ···················· | |
| B | 6218 | hCoV-19/USA/WA-S538/2020\|EPI_ISL_434298\|2020-03-31 | ···················· | ···················· | ···················· | ···················· | ···················· | ···················· | |
| B | 6219 | hCoV-19/USA/WA-S539/2020\|EPI_ISL_434299\|2020-03-23 | ···················· | ···················· | ···················· | ···················· | ···················· | ···················· | |

| | | | | | | | | |
|---|---|---|---|---|---|---|---|---|
| B | 6220 | hCoV-19/USA/NY-NYUMC436/2020\|EPI_ISL_435503\|2020-0 | .................... | .................... | .................... | .................... | .................... | .................... |
| B | 6221 | hCoV-19/USA/NY-NYUMC437/2020\|EPI_ISL_435504\|2020-0 | .................... | .................... | .................... | .................... | .................... | .................... |
| B | 6222 | hCoV-19/USA/NY-NYUMC438/2020\|EPI_ISL_435505\|2020-0 | .................... | .................... | .................... | .................... | .................... | .................... |
| B | 6223 | hCoV-19/USA/NY-NYUMC439/2020\|EPI_ISL_435506\|2020-0 | .................... | .................... | .................... | .................... | .................... | .................... |
| B | 6224 | hCoV-19/USA/NY-NYUMC440/2020\|EPI_ISL_435507\|2020-0 | .................... | .................... | .................... | .................... | .................... | .................... |
| B | 6225 | hCoV-19/USA/NY-NYUMC441/2020\|EPI_ISL_435508\|2020-0 | .................... | .................... | .................... | .................... | .................... | .................... |
| B | 6226 | hCoV-19/USA/NY-NYUMC442/2020\|EPI_ISL_435509\|2020-0 | .................... | .................... | .................... | .................... | .................... | .................... |
| B | 6227 | hCoV-19/USA/NY-NYUMC443/2020\|EPI_ISL_435510\|2020-0 | .................... | .................... | .................... | .................... | .................... | .................... |
| B | 6228 | hCoV-19/USA/NY-NYUMC444/2020\|EPI_ISL_435511\|2020-0 | .................... | .................... | .................... | .................... | .................... | .................... |
| B | 6229 | hCoV-19/USA/NY-NYUMC445/2020\|EPI_ISL_435512\|2020-0 | .................... | .................... | .................... | .................... | .................... | .................... |
| B | 6230 | hCoV-19/USA/NY-NYUMC446/2020\|EPI_ISL_435513\|2020-0 | .................... | .................... | .................... | .................... | .................... | .................... |
| B | 6231 | hCoV-19/USA/NY-NYUMC447/2020\|EPI_ISL_435514\|2020-0 | .................... | .................... | .................... | .................... | .................... | .................... |
| B | 6232 | hCoV-19/USA/NY-NYUMC448/2020\|EPI_ISL_435515\|2020-0 | .................... | .................... | .................... | .................... | .................... | .................... |
| B | 6233 | hCoV-19/USA/NY-NYUMC449/2020\|EPI_ISL_435516\|2020-0 | .................... | .................... | .................... | .................... | .................... | .................... |
| B | 6234 | hCoV-19/USA/NY-NYUMC450/2020\|EPI_ISL_435517\|2020-0 | .................... | .................... | .................... | .................... | .................... | .................... |
| B | 6235 | hCoV-19/USA/NY-NYUMC451/2020\|EPI_ISL_435518\|2020-0 | .................... | .................... | .................... | .................... | .................... | .................... |
| B | 6236 | hCoV-19/USA/NJ-NYUMC452/2020\|EPI_ISL_435519\|2020-0 | .................... | .................... | .................... | .................... | .................... | .................... |
| B | 6237 | hCoV-19/USA/NY-NYUMC453/2020\|EPI_ISL_435520\|2020-0 | .................... | .................... | .................... | .................... | .................... | .................... |
| B | 6238 | hCoV-19/USA/NY-NYUMC455/2020\|EPI_ISL_435521\|2020-0 | .................... | .................... | .................... | .................... | .................... | .................... |
| B | 6239 | hCoV-19/USA/NY-NYUMC456/2020\|EPI_ISL_435522\|2020-0 | .................... | .................... | .................... | .................... | .................... | .................... |
| B | 6240 | hCoV-19/USA/NY-NYUMC457/2020\|EPI_ISL_435523\|2020-0 | .................... | .................... | .................... | .................... | .................... | .................... |
| B | 6241 | hCoV-19/USA/NY-NYUMC458/2020\|EPI_ISL_435524\|2020-0 | .................... | .................... | .................... | .................... | .................... | .................... |
| B | 6242 | hCoV-19/USA/NY-NYUMC459/2020\|EPI_ISL_435525\|2020-0 | .................... | .................... | .................... | .................... | .................... | .................... |
| B | 6243 | hCoV-19/USA/NY-NYUMC460/2020\|EPI_ISL_435526\|2020-0 | .................... | .................... | .................... | .................... | .................... | .................... |
| B | 6244 | hCoV-19/USA/NY-NYUMC461/2020\|EPI_ISL_435527\|2020-0 | .................... | .................... | .................... | .................... | .................... | .................... |
| B | 6245 | hCoV-19/USA/NY-NYUMC462/2020\|EPI_ISL_435528\|2020-0 | .................... | .................... | .................... | .................... | .................... | .................... |
| B | 6246 | hCoV-19/USA/NY-NYUMC464/2020\|EPI_ISL_435529\|2020-0 | .................... | .................... | .................... | .................... | .................... | .................... |
| B | 6247 | hCoV-19/USA/NY-NYUMC465/2020\|EPI_ISL_435530\|2020-0 | .................... | .................... | .................... | .................... | .................... | .................... |
| B | 6248 | hCoV-19/USA/NY-NYUMC466/2020\|EPI_ISL_435531\|2020-0 | .................... | .................... | .................... | .................... | .................... | .................... |
| B | 6249 | hCoV-19/USA/NY-NYUMC467/2020\|EPI_ISL_435532\|2020-0 | .................... | .................... | .................... | .................... | .................... | .................... |
| B | 6250 | hCoV-19/USA/NY-NYUMC468/2020\|EPI_ISL_435533\|2020-0 | .................... | .................... | .................... | .................... | .................... | .................... |
| B | 6251 | hCoV-19/USA/NY-NYUMC469/2020\|EPI_ISL_435534\|2020-0 | .................... | .................... | .................... | .................... | .................... | .................... |
| B | 6252 | hCoV-19/USA/NY-NYUMC470/2020\|EPI_ISL_435535\|2020-0 | .................... | .................... | .................... | .................... | .................... | .................... |
| B | 6253 | hCoV-19/USA/NY-NYUMC471/2020\|EPI_ISL_435536\|2020-0 | .................... | .................... | .................... | .................... | .................... | .................... |
| B | 6254 | hCoV-19/USA/NY-NYUMC472/2020\|EPI_ISL_435537\|2020-0 | .................... | .................... | .................... | .................... | .................... | .................... |
| B | 6255 | hCoV-19/USA/NY-NYUMC473/2020\|EPI_ISL_435538\|2020-0 | .................... | .................... | .................... | .................... | .................... | .................... |
| B | 6256 | hCoV-19/USA/NY-NYUMC474/2020\|EPI_ISL_435539\|2020-0 | .................... | .................... | .................... | .................... | .................... | .................... |
| B | 6257 | hCoV-19/USA/NY-NYUMC475/2020\|EPI_ISL_435540\|2020-0 | .................... | .................... | .................... | .................... | .................... | .................... |
| B | 6258 | hCoV-19/USA/NY-NYUMC476/2020\|EPI_ISL_435541\|2020-0 | .................... | .................... | .................... | .................... | .................... | .................... |
| B | 6259 | hCoV-19/USA/NY-NYUMC477/2020\|EPI_ISL_435542\|2020-0 | .................... | .................... | .................... | .................... | .................... | .................... |
| B | 6260 | hCoV-19/USA/NY-NYUMC478/2020\|EPI_ISL_435543\|2020-0 | .................... | .................... | .................... | .................... | .................... | .................... |
| B | 6261 | hCoV-19/USA/NY-NYUMC479/2020\|EPI_ISL_435544\|2020-0 | .................... | .................... | .................... | .................... | .................... | .................... |
| B | 6262 | hCoV-19/USA/NY-NYUMC480/2020\|EPI_ISL_435545\|2020-0 | .................... | .................... | .................... | .................... | .................... | .................... |
| B | 6263 | hCoV-19/USA/NY-NYUMC481/2020\|EPI_ISL_435546\|2020-0 | .................... | .................... | .................... | .................... | .................... | .................... |
| B | 6264 | hCoV-19/USA/LA-EVTL031/2020\|EPI_ISL_435569\|2020-04 | .................... | .................... | .................... | .................... | .................... | .................... |
| B | 6265 | hCoV-19/USA/LA-EVTL032/2020\|EPI_ISL_435570\|2020-04 | .................... | .................... | .................... | .................... | .................... | .................... |
| B | 6266 | hCoV-19/USA/LA-EVTL034/2020\|EPI_ISL_435572\|2020-04 | .................... | .................... | .................... | .................... | .................... | .................... |
| B | 6267 | hCoV-19/USA/LA-EVTL035/2020\|EPI_ISL_435573\|2020-04 | .................... | .................... | .................... | .................... | .................... | .................... |
| B | 6268 | hCoV-19/USA/LA-EVTL036/2020\|EPI_ISL_435574\|2020-04 | .................... | .................... | .................... | .................... | .................... | .................... |
| B | 6269 | hCoV-19/USA/LA-EVTL037/2020\|EPI_ISL_435575\|2020-04 | .................... | .................... | .................... | .................... | .................... | .................... |
| B | 6270 | hCoV-19/USA/LA-EVTL038/2020\|EPI_ISL_435576\|2020-04 | .................... | .................... | .................... | .................... | .................... | .................... |
| B | 6271 | hCoV-19/USA/LA-EVTL039/2020\|EPI_ISL_435577\|2020-04 | .................... | .................... | .................... | .................... | .................... | .................... |
| B | 6272 | hCoV-19/USA/LA-EVTL040/2020\|EPI_ISL_435578\|2020-04 | .................... | .................... | .................... | .................... | .................... | .................... |
| B | 6273 | hCoV-19/USA/LA-EVTL041/2020\|EPI_ISL_435579\|2020-04 | .................... | .................... | .................... | .................... | .................... | .................... |
| B | 6274 | hCoV-19/USA/CA-SCCPHD-UC115/2020\|EPI_ISL_435594\|20 | .................... | .................... | .................... | .................... | .................... | .................... |
| B | 6275 | hCoV-19/USA/CA-SCCPHD-UC117/2020\|EPI_ISL_435596\|20 | .................... | .................... | .................... | .................... | .................... | .................... |
| B | 6276 | hCoV-19/USA/MI-MDHHS-SC20160/2020\|EPI_ISL_436834\|2 | .................... | .................... | .................... | .................... | .................... | .................... |
| B | 6277 | hCoV-19/USA/MI-MDHHS-SC20106/2020\|EPI_ISL_436836\|2 | .................... | .................... | .................... | .................... | .................... | .................... |
| B | 6278 | hCoV-19/USA/MI-MDHHS-SC20117/2020\|EPI_ISL_436838\|2 | .................... | .................... | .................... | .................... | .................... | .................... |
| B | 6279 | hCoV-19/USA/MI-MDHHS-SC20149/2020\|EPI_ISL_436841\|2 | .................... | .................... | .................... | .................... | .................... | .................... |
| B | 6280 | hCoV-19/USA/MI-MDHHS-SC20152/2020\|EPI_ISL_436842\|2 | .................... | .................... | .................... | .................... | .................... | .................... |
| B | 6281 | hCoV-19/USA/MI-MDHHS-SC20183/2020\|EPI_ISL_436843\|2 | .................... | .................... | .................... | .................... | .................... | .................... |
| B | 6282 | hCoV-19/USA/MI-MDHHS-SC20111/2020\|EPI_ISL_436846\|2 | .................... | .................... | .................... | .................... | .................... | .................... |
| B | 6283 | hCoV-19/USA/MI-MDHHS-SC20116/2020\|EPI_ISL_436847\|2 | .................... | .................... | .................... | .................... | .................... | .................... |
| B | 6284 | hCoV-19/USA/MI-MDHHS-SC20119/2020\|EPI_ISL_436848\|2 | .................... | .................... | .................... | .................... | .................... | .................... |
| B | 6285 | hCoV-19/USA/MI-MDHHS-SC20124/2020\|EPI_ISL_436850\|2 | .................... | .................... | .................... | .................... | .................... | .................... |
| B | 6286 | hCoV-19/USA/MI-MDHHS-SC20134/2020\|EPI_ISL_436852\|2 | .................... | .................... | .................... | .................... | .................... | .................... |
| B | 6287 | hCoV-19/USA/MI-MDHHS-SC20138/2020\|EPI_ISL_436853\|2 | .................... | .................... | .................... | .................... | .................... | .................... |
| B | 6288 | hCoV-19/USA/MI-MDHHS-SC20140/2020\|EPI_ISL_436854\|2 | .................... | .................... | .................... | .................... | .................... | .................... |
| B | 6289 | hCoV-19/USA/MI-MDHHS-SC20142/2020\|EPI_ISL_436855\|2 | .................... | .................... | .................... | .................... | .................... | .................... |
| B | 6290 | hCoV-19/USA/MI-MDHHS-SC20148/2020\|EPI_ISL_436856\|2 | .................... | .................... | .................... | .................... | .................... | .................... |
| B | 6291 | hCoV-19/USA/MI-MDHHS-SC20170/2020\|EPI_ISL_436857\|2 | .................... | .................... | .................... | .................... | .................... | .................... |
| B | 6292 | hCoV-19/USA/MI-MDHHS-SC20180/2020\|EPI_ISL_436859\|2 | .................... | .................... | .................... | .................... | .................... | .................... |
| B | 6293 | hCoV-19/USA/MI-MDHHS-SC20101/2020\|EPI_ISL_436861\|2 | .................... | .................... | .................... | .................... | .................... | .................... |
| B | 6294 | hCoV-19/USA/MI-MDHHS-SC20167/2020\|EPI_ISL_436863\|2 | .................... | .................... | .................... | .................... | .................... | .................... |
| B | 6295 | hCoV-19/USA/MI-MDHHS-SC20174/2020\|EPI_ISL_436864\|2 | .................... | .................... | .................... | .................... | .................... | .................... |
| B | 6296 | hCoV-19/USA/MI-MDHHS-SC20181/2020\|EPI_ISL_436865\|2 | .................... | .................... | .................... | .................... | .................... | .................... |
| B | 6297 | hCoV-19/USA/MI-MDHHS-SC20135/2020\|EPI_ISL_436867\|2 | .................... | .................... | .................... | .................... | .................... | .................... |

| | | | | | | | | |
|---|---|---|---|---|---|---|---|---|
| B | 6298 hCoV-19/USA/MI-MDHHS-SC20157/2020\|EPI_ISL_436868\|2 | ···················· | ···················· | ···················· | ···················· | ···················· | ···················· | |
| B | 6299 hCoV-19/USA/MI-MDHHS-SC20166/2020\|EPI_ISL_436869\|2 | ···················· | ···················· | ···················· | ···················· | ···················· | ···················· | |
| B | 6300 hCoV-19/USA/MI-MDHHS-SC20188/2020\|EPI_ISL_436870\|2 | ···················· | ···················· | ···················· | ···················· | ···················· | ···················· | |
| B | 6301 hCoV-19/USA/MI-MDHHS-SC20144/2020\|EPI_ISL_436874\|2 | ···················· | ···················· | ···················· | ···················· | ···················· | ···················· | |
| B | 6302 hCoV-19/USA/MI-MDHHS-SC20146/2020\|EPI_ISL_436875\|2 | ···················· | ···················· | ···················· | ···················· | ···················· | ···················· | |
| B | 6303 hCoV-19/USA/MI-MDHHS-SC20151/2020\|EPI_ISL_436876\|2 | ···················· | ···················· | ···················· | ···················· | ···················· | ···················· | |
| B | 6304 hCoV-19/USA/MI-MDHHS-SC20155/2020\|EPI_ISL_436877\|2 | ···················· | ···················· | ···················· | ···················· | ···················· | ···················· | |
| B | 6305 hCoV-19/Scotland/CVR809/2020\|EPI_ISL_439116\|2020-0 | ···················· | ···················· | ···················· | ···················· | ···················· | ···················· | |
| B | 6306 hCoV-19/Scotland/CVR817/2020\|EPI_ISL_439117\|2020-0 | ···················· | ···········A········ | ···················· | ···················· | ···················· | ···················· | |
| B | 6307 hCoV-19/Scotland/CVR819/2020\|EPI_ISL_439118\|2020-0 | ···················· | ···················· | ···················· | ···················· | ···················· | ···················· | |
| B | 6308 hCoV-19/Scotland/CVR839/2020\|EPI_ISL_439120\|2020-0 | ···················· | ···················· | ···················· | ···················· | ···················· | ···················· | |
| B | 6309 hCoV-19/Scotland/CVR866/2020\|EPI_ISL_439123\|2020-0 | ···················· | ······A············· | ···················· | ···················· | ···················· | ···················· | |
| B | 6310 hCoV-19/Scotland/CVR871/2020\|EPI_ISL_439125\|2020-0 | ···················· | ···················A | ···················· | ···················· | ···················· | ···················· | 1 |
| B | 6311 hCoV-19/Scotland/CVR876/2020\|EPI_ISL_439126\|2020-0 | ···················· | ···················· | ···················· | ···················· | ···················· | ···················· | |
| B | 6312 hCoV-19/Scotland/CVR878/2020\|EPI_ISL_439127\|2020-0 | ···················· | ···················· | ···················· | ···················· | ···················· | ···················· | |
| B | 6313 hCoV-19/Scotland/CVR978/2020\|EPI_ISL_439140\|2020-0 | ···················· | ···················· | ···················· | ···················· | ···················· | ···················· | |
| B | 6314 hCoV-19/Scotland/CVR989/2020\|EPI_ISL_439142\|2020-0 | ···················· | ···················· | ···················· | ···················· | ···················· | ···················· | |
| B | 6315 hCoV-19/Scotland/EDB1014/2020\|EPI_ISL_439144\|2020- | ···················· | ·A·················· | ···················· | ···················· | ···················· | ···················· | |
| B | 6316 hCoV-19/England/BRIS-121D2F/2020\|EPI_ISL_440100\|20 | ···················· | ···················· | ···················· | ···················· | ···················· | ···················· | |
| B | 6317 hCoV-19/England/BRIS-1222A7/2020\|EPI_ISL_440101\|20 | ···················· | ···················· | ···················· | ···················· | ···················· | ···················· | |
| B | 6318 hCoV-19/England/BRIS-121FB0/2020\|EPI_ISL_440102\|20 | ···················· | ···················· | ···················· | ···················· | ···················· | ···················· | |
| B | 6319 hCoV-19/England/BRIS-121EB3/2020\|EPI_ISL_440103\|20 | ···················· | ···················· | ···················· | ···················· | ···················· | ···················· | |
| B | 6320 hCoV-19/England/CAMB-734C0/2020\|EPI_ISL_440104\|202 | ···················· | ···················· | ···················· | ···················· | ···················· | ···················· | |
| B | 6321 hCoV-19/England/BRIS-121E1D/2020\|EPI_ISL_440105\|20 | ···················· | ···················· | ···················· | ···················· | ···················· | ···················· | |
| B | 6322 hCoV-19/England/BRIS-121DA7/2020\|EPI_ISL_440106\|20 | ···················· | ···················· | ···················· | ···················· | ···················· | ···················· | |
| B | 6323 hCoV-19/England/CAMB-734DF/2020\|EPI_ISL_440107\|202 | ···················· | ···················· | ···················· | ···················· | ···K················ | ···················· | |
| B | 6324 hCoV-19/England/CAMB-72DC6/2020\|EPI_ISL_440108\|202 | ···················· | ···················· | ···················· | ···················· | ···················· | ···················· | |
| B | 6325 hCoV-19/England/CAMB-7333C/2020\|EPI_ISL_440109\|202 | ···················· | ···················· | ···················· | ···················· | ···················· | ···················· | |
| B | 6326 hCoV-19/England/CAMB-73378/2020\|EPI_ISL_440110\|202 | ···················· | ···················· | ···················· | ···················· | ···················· | ···················· | |
| B | 6327 hCoV-19/England/CAMB-735DC/2020\|EPI_ISL_440111\|202 | ···················· | ···················· | ···················· | ···················· | ···················· | ···················· | |
| B | 6328 hCoV-19/England/CAMB-72CAB/2020\|EPI_ISL_440125\|202 | ···················· | ···················· | ···················· | ···················· | ···················· | ···················· | |
| B | 6329 hCoV-19/England/BRIS-1221F5/2020\|EPI_ISL_440126\|20 | ···················· | ···················· | ···················· | ···················· | ···················· | ···················· | |
| B | 6330 hCoV-19/England/BRIS-1222C5/2020\|EPI_ISL_440127\|20 | ···················· | ···················· | ···················· | ···················· | ···················· | ···················· | |
| B | 6331 hCoV-19/England/BRIS-12227A/2020\|EPI_ISL_440128\|20 | ···················· | ···················· | ···················· | ···················· | ···················· | ···················· | |
| B | 6332 hCoV-19/England/CAMB-730CC/2020\|EPI_ISL_440129\|202 | ···················· | ···················· | ···················· | ···················· | ···················· | ···················· | |
| B | 6333 hCoV-19/Northern Ireland/NIRE-FB41B/2020\|EPI_ISL_4 | ···················· | ···················· | ···················· | ···················· | ···················· | ···················· | |
| B | 6334 hCoV-19/Northern Ireland/NIRE-FB42A/2020\|EPI_ISL_4 | ···················· | ···················· | ···················· | ···················· | ···················· | ···················· | |
| B | 6335 hCoV-19/Northern Ireland/NIRE-FB439/2020\|EPI_ISL_4 | ···················· | ···················· | ···················· | ···················· | ···················· | ···················· | |
| B | 6336 hCoV-19/Northern Ireland/NIRE-FB448/2020\|EPI_ISL_4 | ···················· | ···················· | ···················· | ···················· | ···················· | ···················· | |
| B | 6337 hCoV-19/Northern Ireland/NIRE-FB457/2020\|EPI_ISL_4 | ···················· | ···················· | ···················· | ···················· | ···················· | ···················· | |
| B | 6338 hCoV-19/Northern Ireland/NIRE-FB466/2020\|EPI_ISL_4 | ···················· | ···················· | ···················· | ···················· | ···················· | ···················· | |
| B | 6339 hCoV-19/Northern Ireland/NIRE-FB475/2020\|EPI_ISL_4 | ···················· | ···················· | ···················· | ···················· | ···················· | ···················· | |
| B | 6340 hCoV-19/Northern Ireland/NIRE-FB484/2020\|EPI_ISL_4 | ···················· | ···················· | ···················· | ···················· | ···················· | ···················· | |
| B | 6341 hCoV-19/Northern Ireland/NIRE-FB493/2020\|EPI_ISL_4 | ···················· | ···················· | ···················· | ···················· | ···················· | ···················· | |
| B | 6342 hCoV-19/Northern Ireland/NIRE-FB4A2/2020\|EPI_ISL_4 | ···················· | ···················· | ···················· | ···················· | ···················· | ···················· | |
| B | 6343 hCoV-19/Northern Ireland/NIRE-FB4B1/2020\|EPI_ISL_4 | ···················· | ···················· | ···················· | ···················· | ···················· | ···················· | |
| B | 6344 hCoV-19/Northern Ireland/NIRE-FB4C0/2020\|EPI_ISL_4 | ···················· | ···················· | ···················· | ···················· | ···················· | ···················· | |
| B | 6345 hCoV-19/Northern Ireland/NIRE-FB4DF/2020\|EPI_ISL_4 | ···················· | ···················A | ···················· | ···················· | ···················· | ···················· | 1 |
| B | 6346 hCoV-19/Northern Ireland/NIRE-FB4EE/2020\|EPI_ISL_4 | ···················· | ···················· | ···················· | ···················· | ···················· | ···················· | |
| B | 6347 hCoV-19/Northern Ireland/NIRE-FB4FD/2020\|EPI_ISL_4 | ···················· | ···················· | ···················· | ···················· | ···················· | ···················· | |
| B | 6348 hCoV-19/Northern Ireland/NIRE-FB509/2020\|EPI_ISL_4 | ···················· | ···················· | ···················· | ···················· | ···················· | ···················· | |
| B | 6349 hCoV-19/England/BRIS-123BE7/2020\|EPI_ISL_442700\|20 | ···················· | ·A·················· | ···················· | ···················· | ···················· | ···················· | |
| B | 6350 hCoV-19/England/BRIS-12340A/2020\|EPI_ISL_442701\|20 | ···················· | ···················· | ···················· | ···················· | ···················· | ···················· | |
| B | 6351 hCoV-19/England/BRIS-123A9F/2020\|EPI_ISL_442702\|20 | ···················· | ···················· | ···················· | ···················· | ···················· | ···················· | |
| B | 6352 hCoV-19/England/BRIS-123877/2020\|EPI_ISL_442704\|20 | ···················· | ···················· | ···················· | ···················· | ···················· | ···················· | |
| B | 6353 hCoV-19/England/BRIS-1232F1/2020\|EPI_ISL_442705\|20 | ···················· | ···················· | ···················· | ···················· | ···················· | ···················· | |
| B | 6354 hCoV-19/England/BRIS-123B8D/2020\|EPI_ISL_442706\|20 | ···················· | ···················· | ···················· | ···················· | ···················· | ···················· | |
| B | 6355 hCoV-19/England/BRIS-123473/2020\|EPI_ISL_442707\|20 | ···················· | ···················· | ···················· | ···················· | ···················· | ···················· | |
| B | 6356 hCoV-19/England/BRIS-12373E/2020\|EPI_ISL_442708\|20 | ···················· | ···················· | ···················· | ···················· | ···················· | ···················· | |
| B | 6357 hCoV-19/England/BRIS-123701/2020\|EPI_ISL_442709\|20 | ···················· | ···················· | ···················· | ···················· | ···················· | ···················· | |
| B | 6358 hCoV-19/England/BRIS-123C11/2020\|EPI_ISL_442752\|20 | ···················· | ···················· | ···················· | ···················· | ···················· | ···················· | |
| B | 6359 hCoV-19/England/BRIS-1234FB/2020\|EPI_ISL_442753\|20 | ···················· | ···················· | ···················· | ···················· | ···················· | ···················· | |
| B | 6360 hCoV-19/England/BRIS-1232B5/2020\|EPI_ISL_442754\|20 | ···················· | ···················· | ···················· | ···················· | ···················· | ···················· | |
| B | 6361 hCoV-19/England/BRIS-1236F5/2020\|EPI_ISL_442755\|20 | ···················· | ···················· | ···················· | ···················· | ···················· | ···················· | |
| B | 6362 hCoV-19/England/BRIS-123561/2020\|EPI_ISL_442756\|20 | ···················· | ···················· | ···················· | ···················· | ···················· | ···················· | |
| B | 6363 hCoV-19/England/BRIS-123042/2020\|EPI_ISL_442757\|20 | ···················· | ···················· | ···················· | ···················· | ···················· | ···················· | |
| B | 6364 hCoV-19/England/BRIS-12325B/2020\|EPI_ISL_442758\|20 | ···················· | ···················· | ···················· | ···················· | ···················· | ···················· | |
| B | 6365 hCoV-19/England/BRIS-1234DD/2020\|EPI_ISL_442759\|20 | ···················· | ···················· | ···················· | ···················· | ···················· | ···················· | |
| B | 6366 hCoV-19/England/BRIS-123446/2020\|EPI_ISL_442760\|20 | ···················· | ···················· | ···················· | ···················· | ···················· | ···················· | |
| B | 6367 hCoV-19/England/BRIS-12366E/2020\|EPI_ISL_442761\|20 | ···················· | ···················· | ···················· | ···················· | ···················· | ···················· | |
| B | 6368 hCoV-19/England/BRIS-1231A9/2020\|EPI_ISL_442762\|20 | ···················· | ···················· | ···················· | ···················· | ···················· | ···················· | |
| B | 6369 hCoV-19/England/BRIS-12358F/2020\|EPI_ISL_442763\|20 | ···················· | ···················· | ···················· | ···················· | ···················· | ···················· | |
| B | 6370 hCoV-19/England/BRIS-123A35/2020\|EPI_ISL_442764\|20 | ···················· | ···················· | ···················· | ···················· | ···················· | ···················· | |
| B | 6371 hCoV-19/England/BRIS-1237A7/2020\|EPI_ISL_442765\|20 | ···················· | ···················· | ···················· | ···················· | ···················· | ···················· | |
| B | 6372 hCoV-19/England/BRIS-1237F2/2020\|EPI_ISL_442767\|20 | ···················· | ···················· | ···················· | ···················· | ···················· | ···················· | |
| B | 6373 hCoV-19/England/BRIS-123AAE/2020\|EPI_ISL_442768\|20 | ···················· | ···················· | ···················· | ···················· | ···················· | ···················· | |
| B | 6374 hCoV-19/England/BRIS-123B41/2020\|EPI_ISL_442769\|20 | ···················· | ···················· | ···················· | ···················· | ···················· | ···················· | |
| B | 6375 hCoV-19/England/BRIS-123200/2020\|EPI_ISL_442770\|20 | ···················· | ···················· | ···················· | ···················· | ···················· | ···················· | |

| | | | | | | | | |
|---|---|---|---|---|---|---|---|---|
| B | 6376 | hCoV-19/England/BRIS-1239CF/2020\|EPI_ISL_442771\|20 | ···················· | ···················· | ···················· | ···················· | ···················· | ···················· |
| B | 6377 | hCoV-19/England/BRIS-123C6C/2020\|EPI_ISL_442772\|20 | ···················· | ···················· | ···················· | ···················· | ···················· | ···················· |
| B | 6378 | hCoV-19/England/BRIS-1237B6/2020\|EPI_ISL_442773\|20 | ···················· | ·A·················· | ···················· | ···················· | ···················· | ···················· |
| B | 6379 | hCoV-19/England/BRIS-1233EF/2020\|EPI_ISL_442786\|20 | ···················· | ···················· | ···················· | ···················· | ···················· | ···················· |
| B | 6380 | hCoV-19/England/BRIS-123B50/2020\|EPI_ISL_442789\|20 | ···················· | ···················· | ···················· | ···················· | ···················· | ···················· |
| B | 6381 | hCoV-19/Wales/PHWC-2F223/2020\|EPI_ISL_446220\|2020- | ···················· | ···················· | ···················· | ···················· | ···················· | ···················· |
| B | 6382 | hCoV-19/Wales/PHWC-2F232/2020\|EPI_ISL_446221\|2020- | ···················· | ···················· | ···················· | ···················· | ···················· | ···················· |
| B | 6383 | hCoV-19/Wales/PHWC-2F241/2020\|EPI_ISL_446222\|2020- | ···················· | ···················· | ···················· | ···················· | ···················· | ···················· |
| B | 6384 | hCoV-19/Wales/PHWC-2F250/2020\|EPI_ISL_446223\|2020- | ···················· | ···················· | ···················· | ···················· | ···················· | ···················· |
| B | 6385 | hCoV-19/Wales/PHWC-2F26F/2020\|EPI_ISL_446224\|2020- | ···················· | ···················· | ···················· | ···················· | ···················· | ···················· |
| B | 6386 | hCoV-19/Wales/PHWC-2F29C/2020\|EPI_ISL_446227\|2020- | ···················· | ····A··············· | ···················· | ···················· | ···················· | ···················· |
| B | 6387 | hCoV-19/Wales/PHWC-2F37B/2020\|EPI_ISL_446239\|2020- | ···················· | ···················· | ···················· | ···················· | ···················· | ···················· |
| B | 6388 | hCoV-19/Wales/PHWC-2F3A8/2020\|EPI_ISL_446242\|2020- | ···················· | ···················· | ···················· | ···················· | ···················· | ···················· |
| B | 6389 | hCoV-19/Wales/PHWC-2F3B7/2020\|EPI_ISL_446243\|2020- | ···················· | ···················· | ···················· | ···················· | ···················· | ···················· |
| B | 6390 | hCoV-19/Wales/PHWC-2F4B4/2020\|EPI_ISL_446257\|2020- | ···················· | ···················· | ···················· | ···················· | ···················· | ···················· |
| B | 6391 | hCoV-19/Wales/PHWC-2FFFF/2020\|EPI_ISL_446269\|2020- | ···················· | ···················· | ···················· | ···················· | ···················· | ···················· |
| B | 6392 | hCoV-19/Wales/PHWC-30007/2020\|EPI_ISL_446270\|2020- | ···················· | ···················· | ···················· | ···················· | ···················· | ···················· |
| B | 6393 | hCoV-19/Wales/PHWC-3009E/2020\|EPI_ISL_446271\|2020- | ···················· | ···················· | ···················· | ···················· | ···················· | ···················· |
| B | 6394 | hCoV-19/Wales/PHWC-307C6/2020\|EPI_ISL_446280\|2020- | ···················· | ····A··············· | ···················· | ···················· | ···················· | ···················· |
| B | 6395 | hCoV-19/Wales/PHWC-307E4/2020\|EPI_ISL_446282\|2020- | ···················· | ····A··············· | ···················· | ···················· | ···················· | ···················· |
| B | 6396 | hCoV-19/Wales/PHWC-307F3/2020\|EPI_ISL_446283\|2020- | ···················· | ···················· | ···················· | ···················· | ···················· | ···················· |
| B | 6397 | hCoV-19/Wales/PHWC-3081E/2020\|EPI_ISL_446285\|2020- | ···················· | ···················· | ···················· | ···················· | ···················· | ···················· |
| B | 6398 | hCoV-19/Wales/PHWC-3083C/2020\|EPI_ISL_446287\|2020- | ···················· | ···················· | ···················· | ···················· | ···················· | ···················· |
| B | 6399 | hCoV-19/Wales/PHWC-308C3/2020\|EPI_ISL_446288\|2020- | ···················· | ···················· | ···················· | ···················· | ···················· | ···················· |
| B | 6400 | hCoV-19/Wales/PHWC-3092A/2020\|EPI_ISL_446294\|2020- | ···················· | ···················· | ···················· | ···················· | ···················· | ···················· |
| B | 6401 | hCoV-19/USA/WA-UW-5182/2020\|EPI_ISL_430872\|2020-04 | ···················· | ···················· | ···················· | ···················· | ···················· | ···················· |
| B | 6402 | hCoV-19/USA/WA-UW-5189/2020\|EPI_ISL_430873\|2020-04 | ···················· | ···················· | ···················· | ···················· | ···················· | ···················· |
| B | 6403 | hCoV-19/USA/WA-UW-5190/2020\|EPI_ISL_430874\|2020-04 | ···················· | ···················· | ···················· | ···················· | ···················· | ···················· |
| B | 6404 | hCoV-19/USA/WA-UW-524/2020\|EPI_ISL_430877\|2020-03- | ···················· | ···················· | ···················· | ···················· | ···················· | ···················· |
| B | 6405 | hCoV-19/USA/WA-UW-525/2020\|EPI_ISL_430878\|2020-03- | ···················· | ···················· | ···················· | ···················· | ···················· | ···················· |
| B | 6406 | hCoV-19/USA/UN-UW-526/2020\|EPI_ISL_430879\|2020-03- | ···················· | ···················· | ···················· | ···················· | ···················· | ···················· |
| B | 6407 | hCoV-19/USA/WA-UW-530/2020\|EPI_ISL_430880\|2020-03- | ···················· | ···················· | ···················· | ···················· | ···················· | ···················· |
| B | 6408 | hCoV-19/USA/UN-UW-531/2020\|EPI_ISL_430881\|2020-03- | ···················· | ···················· | ···················· | ···················· | ···················· | ···················· |
| B | 6409 | hCoV-19/USA/UN-UW-536/2020\|EPI_ISL_430882\|2020-03- | ···················· | ···················· | ···················· | ···················· | ···················· | ···················· |
| B | 6410 | hCoV-19/USA/WA-UW-542/2020\|EPI_ISL_430883\|2020-03- | ···················· | ···················· | ···················· | ···················· | ···················· | ···················· |
| B | 6411 | hCoV-19/USA/WA-UW-543/2020\|EPI_ISL_430884\|2020-03- | ···················· | ···················· | ···················· | ···················· | ···················· | ···················· |
| B | 6412 | hCoV-19/USA/WA-UW-546/2020\|EPI_ISL_430885\|2020-03- | ···················· | ···················· | ···················· | ···················· | ···················· | ···················· |
| B | 6413 | hCoV-19/USA/UN-UW-547/2020\|EPI_ISL_430886\|2020-03- | ···················· | ···················· | ···················· | ···················· | ···················· | ···················· |
| B | 6414 | hCoV-19/USA/WA-UW-555/2020\|EPI_ISL_430887\|2020-03- | ···················· | ···················· | ···················· | ···················· | ···················· | ···················· |
| B | 6415 | hCoV-19/USA/WA-UW-558/2020\|EPI_ISL_430888\|2020-03- | ···················· | ···················· | ···················· | ···················· | ···················· | ···················· |
| B | 6416 | hCoV-19/USA/WA-UW-572/2020\|EPI_ISL_430889\|2020-03- | ···················· | ···················· | ···················· | ···················· | ···················· | ···················· |
| B | 6417 | hCoV-19/USA/UN-UW-581/2020\|EPI_ISL_430890\|2020-03- | ···················· | ···················· | ···················· | ···················· | ···················· | ···················· |
| B | 6418 | hCoV-19/USA/WA-UW-586/2020\|EPI_ISL_430891\|2020-03- | ···················· | ···················· | ···················· | ···················· | ···················· | ···················· |
| B | 6419 | hCoV-19/USA/WA-UW-596/2020\|EPI_ISL_430892\|2020-03- | ···················· | ···················· | ···················· | ···················· | ···················· | ···················· |
| B | 6420 | hCoV-19/USA/WA-UW-600/2020\|EPI_ISL_430893\|2020-03- | ···················· | ···················· | ···················· | ···················· | ···················· | ···················· |
| B | 6421 | hCoV-19/USA/WA-S456/2020\|EPI_ISL_434216\|2020-04-06 | ···················· | ···················· | ···················· | ···················· | ···················· | ···················· |
| B | 6422 | hCoV-19/USA/WA-S457/2020\|EPI_ISL_434217\|2020-04-02 | ···················· | ···················· | ···················· | ···················· | ···················· | ···················· |
| B | 6423 | hCoV-19/USA/WA-S458/2020\|EPI_ISL_434218\|2020-04-06 | ···················· | ···················· | ···················· | ···················· | ···················· | ···················· |
| B | 6424 | hCoV-19/USA/WA-S459/2020\|EPI_ISL_434219\|2020-04-06 | ···················· | ···················· | ···················· | ···················· | ···················· | ···················· |
| B | 6425 | hCoV-19/USA/WA-S460/2020\|EPI_ISL_434220\|2020-04-05 | ···················· | ···················· | ···················· | ···················· | ···················· | ···················· |
| B | 6426 | hCoV-19/USA/WA-S461/2020\|EPI_ISL_434221\|2020-04-06 | ···················· | ···················· | ···················· | ···················· | ···················· | ···················· |
| B | 6427 | hCoV-19/USA/WA-S462/2020\|EPI_ISL_434222\|2020-04-02 | ···················· | ···················· | ···················· | ···················· | ···················· | ···················· |
| B | 6428 | hCoV-19/USA/WA-S463/2020\|EPI_ISL_434223\|2020-04-04 | ···················· | ···················· | ···················· | ···················· | ···················· | ···················· |
| B | 6429 | hCoV-19/USA/WA-S464/2020\|EPI_ISL_434224\|2020-04-04 | ···················· | ···················· | ···················· | ···················· | ···················· | ···················· |
| B | 6430 | hCoV-19/USA/WA-S465/2020\|EPI_ISL_434225\|2020-04-01 | ···················· | ···················· | ···················· | ···················· | ···················· | ···················· |
| B | 6431 | hCoV-19/USA/WA-S466/2020\|EPI_ISL_434226\|2020-04-01 | ···················· | ···················· | ···················· | ···················· | ···················· | ···················· |
| B | 6432 | hCoV-19/USA/WA-S467/2020\|EPI_ISL_434227\|2020-04-02 | ···················· | ···················· | ···················· | ···················· | ···················· | ···················· |
| B | 6433 | hCoV-19/USA/WA-S468/2020\|EPI_ISL_434228\|2020-04-05 | ···················· | ···················· | ···················· | ···················· | ···················· | ···················· |
| B | 6434 | hCoV-19/USA/WA-S469/2020\|EPI_ISL_434229\|2020-04-05 | ···················· | ···················· | ···················· | ···················· | ···················· | ···················· |
| B | 6435 | hCoV-19/USA/WA-S470/2020\|EPI_ISL_434230\|2020-04-05 | ···················· | ···················· | ···················· | ···················· | ···················· | ···················· |
| B | 6436 | hCoV-19/USA/WA-S471/2020\|EPI_ISL_434231\|2020-04-06 | ···················· | ···················· | ···················· | ···················· | ···················· | ···················· |
| B | 6437 | hCoV-19/USA/WA-S472/2020\|EPI_ISL_434232\|2020-04-05 | ···················· | ···················· | ···················· | ···················· | ···················· | ···················· |
| B | 6438 | hCoV-19/USA/WA-S473/2020\|EPI_ISL_434233\|2020-04-03 | ···················· | ···················· | ···················· | ···················· | ···················· | ···················· |
| B | 6439 | hCoV-19/USA/WA-S474/2020\|EPI_ISL_434234\|2020-04-01 | ···················· | ···················· | ···················· | ···················· | ···················· | ···················· |
| B | 6440 | hCoV-19/USA/WA-S475/2020\|EPI_ISL_434235\|2020-04-01 | ···················· | ···················· | ···················· | ···················· | ···················· | ···················· |
| B | 6441 | hCoV-19/USA/WA-S476/2020\|EPI_ISL_434236\|2020-04-03 | ···················· | ···················· | ···················· | ···················· | ···················· | ···················· |
| B | 6442 | hCoV-19/USA/WA-S477/2020\|EPI_ISL_434237\|2020-04-05 | ···················· | ···················· | ···················· | ···················· | ···················· | ···················· |
| B | 6443 | hCoV-19/USA/WA-S480/2020\|EPI_ISL_434240\|2020-04-05 | ···················· | ···················· | ···················· | ···················· | ···················· | ···················· |
| B | 6444 | hCoV-19/USA/WA-S489/2020\|EPI_ISL_434249\|2020-04-02 | ···················· | ···················· | ···················· | ···················· | ···················· | ···················· |
| B | 6445 | hCoV-19/USA/WA-S490/2020\|EPI_ISL_434250\|2020-03-31 | ···················· | ···················· | ···················· | ···················· | ···················· | ···················· |
| B | 6446 | hCoV-19/USA/WA-S491/2020\|EPI_ISL_434251\|2020-04-02 | ···················· | ···················· | ···················· | ···················· | ···················· | ···················· |
| B | 6447 | hCoV-19/USA/WA-S492/2020\|EPI_ISL_434252\|2020-03-26 | ···················· | ···················· | ···················· | ···················· | ···················· | ···················· |
| B | 6448 | hCoV-19/USA/WA-S493/2020\|EPI_ISL_434253\|2020-04-01 | ···················· | ···················· | ···················· | ···················· | ···················· | ···················· |
| B | 6449 | hCoV-19/USA/WA-S494/2020\|EPI_ISL_434254\|2020-04-01 | ···················· | ···················· | ···················· | ···················· | ···················· | ···················· |
| B | 6450 | hCoV-19/USA/WA-S495/2020\|EPI_ISL_434255\|2020-04-01 | ···················· | ···················· | ···················· | ···················· | ···················· | ···················· |
| B | 6451 | hCoV-19/USA/WA-S496/2020\|EPI_ISL_434256\|2020-03-31 | ···················· | ···················· | ···················· | ···················· | ···················· | ···················· |
| B | 6452 | hCoV-19/USA/WA-S497/2020\|EPI_ISL_434257\|2020-03-31 | ···················· | ···················· | ···················· | ···················· | ···················· | ···················· |
| B | 6453 | hCoV-19/USA/WA-S498/2020\|EPI_ISL_434258\|2020-04-01 | ···················· | ···················· | ···················· | ···················· | ···················· | ···················· |

| | | | | | | | | | |
|---|---|---|---|---|---|---|---|---|---|
| B | 6454 | hCoV-19/USA/WA-S499/2020\|EPI_ISL_434259\|2020-03-31 | ···················· | ···················· | ···················· | ···················· | ···················· | ···················· | |
| B | 6455 | hCoV-19/USA/NY-NYUMC482/2020\|EPI_ISL_435547\|2020-0 | ···················· | ···················· | ···················· | ···················· | ···················· | ···················· | |
| B | 6456 | hCoV-19/USA/NY-NYUMC483/2020\|EPI_ISL_435548\|2020-0 | ···················· | ···················· | ···················· | ···················· | ···················· | ···················· | |
| B | 6457 | hCoV-19/USA/NY-NYUMC484/2020\|EPI_ISL_435549\|2020-0 | ···················· | ···················· | ···················· | ···················· | ···················· | ···················· | |
| B | 6458 | hCoV-19/USA/LA-EVTL012/2020\|EPI_ISL_435550\|2020-04 | ···················· | ···················· | ···················· | ···················· | ···················· | ···················· | |
| B | 6459 | hCoV-19/USA/LA-EVTL013/2020\|EPI_ISL_435551\|2020-04 | ···················· | ···················· | ···················· | ···················· | ···················· | ···················· | |
| B | 6460 | hCoV-19/USA/LA-EVTL014/2020\|EPI_ISL_435552\|2020-04 | ···················· | ···················· | ···················· | ···················· | ···················· | ···················· | |
| B | 6461 | hCoV-19/USA/LA-EVTL015/2020\|EPI_ISL_435553\|2020-04 | ···················· | ···················· | ···················· | ···················· | ···················· | ···················· | |
| B | 6462 | hCoV-19/USA/LA-EVTL016/2020\|EPI_ISL_435554\|2020-04 | ···················· | ···················· | ···················· | ···················· | ···················· | ···················· | |
| B | 6463 | hCoV-19/USA/LA-EVTL017/2020\|EPI_ISL_435555\|2020-04 | ···················· | ···················· | ···················· | ···················· | ···················· | ···················· | |
| B | 6464 | hCoV-19/USA/LA-EVTL018/2020\|EPI_ISL_435556\|2020-04 | ···················· | ···················· | ···················· | ···················· | ···················· | ···················· | |
| B | 6465 | hCoV-19/USA/LA-EVTL019/2020\|EPI_ISL_435557\|2020-04 | ···················· | ···················· | ···················· | ···················· | ···················· | ···················· | |
| B | 6466 | hCoV-19/USA/LA-EVTL020/2020\|EPI_ISL_435558\|2020-04 | ···················· | ···················· | ···················· | ···················· | ···················· | ···················· | |
| B | 6467 | hCoV-19/USA/LA-EVTL021/2020\|EPI_ISL_435559\|2020-04 | ···················· | ···················· | ···················· | ···················· | ···················· | ···················· | |
| B | 6468 | hCoV-19/USA/LA-EVTL022/2020\|EPI_ISL_435560\|2020-04 | ···················· | ···················· | ···················· | ···················· | ···················· | ···················· | |
| B | 6469 | hCoV-19/USA/LA-EVTL023/2020\|EPI_ISL_435561\|2020-04 | ···················· | ···················· | ···················· | ···················· | ···················· | ···················· | |
| B | 6470 | hCoV-19/USA/LA-EVTL024/2020\|EPI_ISL_435562\|2020-04 | ···················· | ···················· | ···················· | ···················· | ···················· | ···················· | |
| B | 6471 | hCoV-19/USA/LA-EVTL025/2020\|EPI_ISL_435563\|2020-04 | ···················· | ···················· | ···················· | ···················· | ···················· | ···················· | |
| B | 6472 | hCoV-19/USA/LA-EVTL026/2020\|EPI_ISL_435564\|2020-04 | ···················· | ···················· | ···················· | ···················· | ···················· | ···················· | |
| B | 6473 | hCoV-19/USA/LA-EVTL027/2020\|EPI_ISL_435565\|2020-04 | ···················· | ···················· | ···················· | ···················· | ···················· | ···················· | |
| B | 6474 | hCoV-19/USA/LA-EVTL028/2020\|EPI_ISL_435566\|2020-04 | ···················· | ···················· | ···················· | ···················· | ···················· | ···················· | |
| B | 6475 | hCoV-19/USA/LA-EVTL029/2020\|EPI_ISL_435567\|2020-04 | ···················· | ···················· | ···················· | ···················· | ···················· | ···················· | |
| B | 6476 | hCoV-19/USA/LA-EVTL030/2020\|EPI_ISL_435568\|2020-04 | ···················· | ···················· | ···················· | ···················· | ···················· | ···················· | |
| B | 6477 | hCoV-19/USA/LA-EVTL033/2020\|EPI_ISL_435571\|2020-04 | ···················· | ···················· | ···················· | ···················· | ···················· | ···················· | |
| B | 6478 | hCoV-19/USA/CA-SCCPHD-UC101/2020\|EPI_ISL_435580\|20 | ···················· | ···················· | ···················· | ···················· | ···················· | ···················· | |
| B | 6479 | hCoV-19/USA/CA-SCCPHD-UC104/2020\|EPI_ISL_435583\|20 | ···················· | ···················· | ···················· | ···················· | ···················· | ···················· | |
| B | 6480 | hCoV-19/USA/CA-SCCPHD-UC108/2020\|EPI_ISL_435587\|20 | ···················· | ···················· | ···················· | ···················· | ···················· | ···················· | |
| B | 6481 | hCoV-19/Singapore/95/2020\|EPI_ISL_435700\|2020-03-1 | ···················· | ···················· | ···················· | ···················· | ···················· | ···················· | |
| B | 6482 | hCoV-19/USA/CT-Yale-100/2020\|EPI_ISL_435702\|2020-0 | ···················· | ···················· | ···················· | ···················· | ···················· | ···················· | |
| B | 6483 | hCoV-19/USA/CT-Yale-105/2020\|EPI_ISL_435703\|2020-0 | ···················· | ···················· | ···················· | ···················· | ···················· | ···················· | |
| B | 6484 | hCoV-19/USA/CT-Yale-106/2020\|EPI_ISL_435704\|2020-0 | ···················· | ···················· | ···················· | ···················· | ···················· | ···················· | |
| B | 6485 | hCoV-19/USA/CT-Yale-108/2020\|EPI_ISL_435705\|2020-0 | ···················· | ···················· | ···················· | ···················· | ···················· | ···················· | |
| B | 6486 | hCoV-19/USA/CT-Yale-109/2020\|EPI_ISL_435706\|2020-0 | ···················· | ···················· | ···················· | ···················· | ···················· | ···················· | |
| B | 6487 | hCoV-19/USA/CT-Yale-112/2020\|EPI_ISL_435708\|2020-0 | ···················· | ···················· | ···················· | ···················· | ···················· | ···················· | |
| B | 6488 | hCoV-19/USA/CT-Yale-139/2020\|EPI_ISL_435709\|2020-0 | ···················· | ···················· | ···················· | ···················· | ···················· | ···················· | |
| B | 6489 | hCoV-19/USA/CT-Yale-146/2020\|EPI_ISL_435716\|2020-0 | ···················· | ···················· | ···················· | ···················· | ···················· | ···················· | |
| B | 6490 | hCoV-19/USA/CT-Yale-151/2020\|EPI_ISL_435720\|2020-0 | ···················· | ···················· | ···················· | ···················· | ···················· | ···················· | |
| B | 6491 | hCoV-19/USA/MI-MDHHS-SC20165/2020\|EPI_ISL_436878\|2 | ···················· | ···················· | ···················· | ···················· | ···················· | ···················· | |
| B | 6492 | hCoV-19/USA/MI-MDHHS-SC20168/2020\|EPI_ISL_436879\|2 | ···················· | ···················· | ···················· | ···················· | ···················· | ···················· | |
| B | 6493 | hCoV-19/USA/MI-MDHHS-SC20178/2020\|EPI_ISL_436880\|2 | ···················· | ···················· | ···················· | ···················· | ···················· | ···················· | |
| B | 6494 | hCoV-19/USA/MI-MDHHS-SC20104/2020\|EPI_ISL_436884\|2 | ···················· | ···················· | ···················· | ···················· | ···················· | ···················· | |
| B | 6495 | hCoV-19/USA/MI-MDHHS-SC20109/2020\|EPI_ISL_436885\|2 | ···················· | ···················· | ···················· | ···················· | ···················· | ···················· | |
| B | 6496 | hCoV-19/USA/MI-MDHHS-SC20115/2020\|EPI_ISL_436886\|2 | ···················· | ···················· | ···················· | ···················· | ···················· | ···················· | |
| B | 6497 | hCoV-19/USA/MI-MDHHS-SC20164/2020\|EPI_ISL_436887\|2 | ···················· | ···················· | ···················· | ···················· | ···················· | ···················· | |
| B | 6498 | hCoV-19/USA/MI-MDHHS-SC20173/2020\|EPI_ISL_436889\|2 | ···················· | ···················· | ···················· | ···················· | ···················· | ···················· | |
| B | 6499 | hCoV-19/USA/WI-GMF-01978/2020\|EPI_ISL_436892\|2020- | ···················· | ···················· | ···················· | ···················· | ·········T·········· | ···················· | |
| B | 6500 | hCoV-19/USA/WI-GMF-01992/2020\|EPI_ISL_436893\|2020- | ···················· | ···················· | ···················· | ···················· | ···················· | ···················· | |
| B | 6501 | hCoV-19/USA/WI-GMF-01993/2020\|EPI_ISL_436894\|2020- | ···················· | ···················· | ···················· | ···················· | ···················· | ···················· | |
| B | 6502 | hCoV-19/USA/WI-GMF-01999/2020\|EPI_ISL_436895\|2020- | ···················· | ···················· | ···················· | ···················· | ···················· | ···················· | |
| B | 6503 | hCoV-19/USA/WI-GMF-02263/2020\|EPI_ISL_436896\|2020- | ···················· | ···················· | ···················· | ···················· | ·········T·········· | ···················· | |
| B | 6504 | hCoV-19/USA/WI-GMF-02274/2020\|EPI_ISL_436897\|2020- | ···················· | ···················· | ···················· | ···················· | ···················· | ···················· | |
| B | 6505 | hCoV-19/USA/WI-GMF-02294/2020\|EPI_ISL_436898\|2020- | ···················· | ···················· | ···················· | ···················· | ·········T·········· | ···················· | |
| B | 6506 | hCoV-19/USA/WI-GMF-02314/2020\|EPI_ISL_436899\|2020- | ···················· | ···················· | ···················· | ···················· | ···················· | ···················· | |
| B | 6507 | hCoV-19/Scotland/EDB2446/2020\|EPI_ISL_439210\|2020- | ···················· | ···················· | ···················· | ···················· | ···················· | ···················· | |
| B | 6508 | hCoV-19/Scotland/EDB2451/2020\|EPI_ISL_439211\|2020- | ···················· | ···················· | ···················· | ···················· | ···················· | ···················· | |
| B | 6509 | hCoV-19/Scotland/EDB2458/2020\|EPI_ISL_439213\|2020- | ···················· | ·A·················· | ···················· | ···················· | ···················· | ···················· | |
| B | 6510 | hCoV-19/Scotland/EDB2462/2020\|EPI_ISL_439214\|2020- | ···················· | ···················· | ···················· | ···················· | ···················· | ···················· | |
| B | 6511 | hCoV-19/Scotland/EDB2463/2020\|EPI_ISL_439215\|2020- | ···················· | ···················· | ···················· | ···················· | ···················· | ···················· | |
| B | 6512 | hCoV-19/Scotland/EDB2466/2020\|EPI_ISL_439216\|2020- | ···················· | ···················· | ···················· | ···················· | ···················· | ···················· | |
| B | 6513 | hCoV-19/Scotland/EDB2467/2020\|EPI_ISL_439217\|2020- | ···················· | ···················· | ···················· | ···················· | ···················· | ···················· | |
| B | 6514 | hCoV-19/Scotland/EDB2468/2020\|EPI_ISL_439218\|2020- | ···················· | ···················· | ···················· | ···················· | ···················· | ···················· | |
| B | 6515 | hCoV-19/Scotland/EDB2471/2020\|EPI_ISL_439219\|2020- | ···················· | ·A·················· | ···················· | ···················· | ···················· | ···················· | |
| B | 6516 | hCoV-19/Scotland/EDB2488/2020\|EPI_ISL_439222\|2020- | ···················· | ···················· | ···················· | ···················· | ···················· | ···················· | |
| B | 6517 | hCoV-19/Scotland/EDB2518/2020\|EPI_ISL_439223\|2020- | ···················· | ···················· | ···················· | ···················· | ···················· | ···················· | |
| B | 6518 | hCoV-19/Scotland/EDB2533/2020\|EPI_ISL_439224\|2020- | ···················· | ···················· | ···················· | ···················· | ···················· | ···················· | |
| B | 6519 | hCoV-19/Scotland/EDB2534/2020\|EPI_ISL_439225\|2020- | ···················· | ·A·················· | ···················· | ···················· | ···················· | ···················· | |
| B | 6520 | hCoV-19/Scotland/EDB2540/2020\|EPI_ISL_439226\|2020- | ···················· | ···················· | ···················· | ···················· | ···················· | ···················· | |
| B | 6521 | hCoV-19/Scotland/EDB2543/2020\|EPI_ISL_439227\|2020- | ···················· | ·A·················· | ···················· | ···················· | ···················· | ···················· | |
| B | 6522 | hCoV-19/Scotland/EDB2554/2020\|EPI_ISL_439228\|2020- | ···················· | ···················· | ···················· | ···················· | ···················· | ···················· | |
| B | 6523 | hCoV-19/Scotland/EDB2557/2020\|EPI_ISL_439229\|2020- | ···················· | ···················· | ···················· | ···················· | ···················· | ···················· | |
| B | 6524 | hCoV-19/Scotland/EDB3497/2020\|EPI_ISL_439270\|2020- | ···················· | ···················A | ···················· | ···················· | ···················· | ···················· | 1 |
| B | 6525 | hCoV-19/Scotland/EDB3498/2020\|EPI_ISL_439271\|2020- | ···················· | ···················A | ···················· | ···················· | ···················· | ···················· | 1 |
| B | 6527 | hCoV-19/Scotland/EDB3500/2020\|EPI_ISL_439273\|2020- | ···················· | ···················A | ···················· | ···················· | ···················· | ···················· | 1 |
| B | 6528 | hCoV-19/Scotland/EDB3501/2020\|EPI_ISL_439274\|2020- | ···················· | ···················· | ···················· | ···················· | ···················· | ···················· | |
| B | 6529 | hCoV-19/Scotland/EDB3502/2020\|EPI_ISL_439275\|2020- | ···················· | ···················A | ···················· | ···················· | ···················· | ···················· | 1 |
| B | 6530 | hCoV-19/Scotland/EDB3503/2020\|EPI_ISL_439276\|2020- | ···················· | ···················A | ···················· | ···················· | ···················· | ···················· | 1 |
| B | 6531 | hCoV-19/Scotland/EDB3518/2020\|EPI_ISL_439277\|2020- | ···················· | ···················· | ···················· | ···················· | ···················· | ···················· | |
| B | 6532 | hCoV-19/Scotland/EDB3520/2020\|EPI_ISL_439278\|2020- | ···················· | ·A·················· | ···················· | ···················· | ···················· | ···················· | |

| | | | | | | | | |
|---|---|---|---|---|---|---|---|---|
| B | 6533 hCoV-19/Scotland/EDB3530/2020\|EPI_ISL_439279\|2020- | ···················· | ···················A | ···················· | ···················· | ···················· | ···················· | 1 |
| B | 6534 hCoV-19/Scotland/EDB3531/2020\|EPI_ISL_439280\|2020- | ···················· | ···················· | ···················· | ···················· | ···················· | ···················· | |
| B | 6535 hCoV-19/Scotland/EDB3532/2020\|EPI_ISL_439281\|2020- | ···················· | ·A·················· | ···················· | ···················· | ···················· | ···················· | |
| B | 6536 hCoV-19/Scotland/EDB3554/2020\|EPI_ISL_439282\|2020- | ···················· | ···················A | ···················· | ···················· | ···················· | ···················· | 1 |
| B | 6537 hCoV-19/Scotland/EDB3555/2020\|EPI_ISL_439283\|2020- | ···················· | ·A·················· | ···················· | ···················· | ···················· | ···················· | |
| B | 6538 hCoV-19/Scotland/EDB3557/2020\|EPI_ISL_439284\|2020- | ···················· | ···················· | ···················· | ···················· | ···················· | ···················· | |
| B | 6539 hCoV-19/Scotland/EDB3558/2020\|EPI_ISL_439285\|2020- | ···················· | ···················· | ···················· | ···················· | ···················· | ···················· | |
| B | 6540 hCoV-19/Scotland/EDB3559/2020\|EPI_ISL_439286\|2020- | ···················· | ···················· | ···················· | ···················· | ···················· | ···················· | |
| B | 6541 hCoV-19/Scotland/EDB3566/2020\|EPI_ISL_439287\|2020- | ···················· | ···················A | ···················· | ···················· | ···················· | ···················· | 1 |
| B | 6542 hCoV-19/Scotland/EDB3568/2020\|EPI_ISL_439288\|2020- | ···················· | ···················· | ···················· | ···················· | ···················· | ···················· | |
| B | 6544 hCoV-19/Scotland/EDB3573/2020\|EPI_ISL_439290\|2020- | ···················· | ···················· | ···················· | ···················· | ···················· | ···················· | |
| B | 6545 hCoV-19/Scotland/EDB3574/2020\|EPI_ISL_439291\|2020- | ···················· | ···················· | ···················· | ···················· | ···················· | ···················· | |
| B | 6546 hCoV-19/Scotland/EDB3576/2020\|EPI_ISL_439292\|2020- | ···················· | ···················· | ···················· | ···················· | ···················· | ···················· | |
| B | 6547 hCoV-19/Scotland/EDB3583/2020\|EPI_ISL_439293\|2020- | ···················· | ···················· | ···················· | ···················· | ···················· | ···················· | |
| B | 6548 hCoV-19/Scotland/EDB3588/2020\|EPI_ISL_439294\|2020- | ···················· | ···················A | ···················· | ···················· | ···················· | ···················· | 1 |
| B | 6549 hCoV-19/Scotland/EDB3598/2020\|EPI_ISL_439296\|2020- | ···················· | ···················· | ···················· | ···················· | ···················· | ···················· | |
| B | 6550 hCoV-19/Scotland/EDB3600/2020\|EPI_ISL_439297\|2020- | ···················· | ···················· | ···················· | ···················· | ···················· | ···················· | |
| B | 6552 hCoV-19/Scotland/EDB3627/2020\|EPI_ISL_439299\|2020- | ···················· | ···················· | ···················· | ···················· | ···················· | ···················· | |
| B | 6553 hCoV-19/England/BRIS-121ABF/2020\|EPI_ISL_440201\|20 | ···················· | ···················· | ···················· | ···················· | ···················· | ···················· | |
| B | 6554 hCoV-19/England/CAMB-73071/2020\|EPI_ISL_440208\|202 | ···················· | ···················· | ···················· | ···················· | ···················· | ···················· | |
| B | 6555 hCoV-19/England/BRIS-121B9E/2020\|EPI_ISL_440255\|20 | ···T················ | ···················· | ···················· | ···················· | ···················· | ···················· | |
| B | 6556 hCoV-19/England/BRIS-12177C/2020\|EPI_ISL_440256\|20 | ···················· | ···················· | ···················· | ···················· | ···················· | ···················· | |
| B | 6557 hCoV-19/England/CAMB-7330F/2020\|EPI_ISL_440257\|202 | ···················· | ···················· | ···················· | ···················· | ···················· | ···················· | |
| B | 6558 hCoV-19/England/CAMB-734FD/2020\|EPI_ISL_440258\|202 | ···················· | ···················· | ···················· | ···················· | ···················· | ···················· | |
| B | 6559 hCoV-19/England/CAMB-733A5/2020\|EPI_ISL_440259\|202 | ···················· | ···················· | ···················· | ···················· | ···················· | ···················· | |
| B | 6560 hCoV-19/England/CAMB-73606/2020\|EPI_ISL_440260\|202 | ···················· | ···················· | ···················· | ···················· | ···················· | ···················· | |
| B | 6561 hCoV-19/England/CAMB-7368E/2020\|EPI_ISL_440261\|202 | ···················· | ···················· | ···················· | ···················· | ···················· | ···················· | |
| B | 6562 hCoV-19/England/CAMB-7317E/2020\|EPI_ISL_440264\|202 | ···················· | ···················· | ···················· | ···················· | ···················· | ···················· | |
| B | 6563 hCoV-19/England/CAMB-731BA/2020\|EPI_ISL_440265\|202 | ···················· | ···················· | ···················· | ···················· | ···················· | ···················· | |
| B | 6564 hCoV-19/England/CAMB-7328A/2020\|EPI_ISL_440267\|202 | ···················· | ···················· | ···················· | ···················· | ···················· | ···················· | |
| B | 6565 hCoV-19/England/CAMB-7331E/2020\|EPI_ISL_440268\|202 | ···················· | ···················· | ···················· | ···················· | ···················· | ···················· | |
| B | 6566 hCoV-19/England/BRIS-121A64/2020\|EPI_ISL_440269\|20 | ···················· | ···················· | ···················· | ···················· | ···················· | ···················· | |
| B | 6567 hCoV-19/England/CAMB-732A8/2020\|EPI_ISL_440270\|202 | ···T················ | ···················· | ···················· | ···················· | ···················· | ···················· | |
| B | 6568 hCoV-19/England/CAMB-7319C/2020\|EPI_ISL_440271\|202 | ···················· | ···················· | ···················· | ···················· | ···················· | ···················· | |
| B | 6569 hCoV-19/England/BRIS-121A46/2020\|EPI_ISL_440272\|20 | ···················· | ···················· | ···················· | ···················· | ···················· | ···················· | |
| B | 6570 hCoV-19/England/BRIS-12192B/2020\|EPI_ISL_440275\|20 | ···················· | ···················· | ···················· | ···················· | ···················· | ···················· | |
| B | 6571 hCoV-19/England/CAMB-7326C/2020\|EPI_ISL_440278\|202 | ···················· | ···················· | ···················· | ···················· | ···················· | ···················· | |
| B | 6572 hCoV-19/England/CAMB-72FC0/2020\|EPI_ISL_440279\|202 | ···················· | ···················· | ···················· | ···················· | ···················· | ···················· | |
| B | 6573 hCoV-19/England/CAMB-73493/2020\|EPI_ISL_440280\|202 | ···T················ | ···················· | ···················· | ···················· | ···················· | ···················· | |
| B | 6574 hCoV-19/England/CAMB-731F6/2020\|EPI_ISL_440282\|202 | ···················· | ···················· | ···················· | ···················· | ···················· | ···················· | |
| B | 6575 hCoV-19/England/CAMB-7318D/2020\|EPI_ISL_440283\|202 | ···················· | ···················· | ···················· | ···················· | ···················· | ···················· | |
| B | 6576 hCoV-19/England/CAMB-73572/2020\|EPI_ISL_440284\|202 | ···················· | ···················· | ···················· | ···················· | ···················· | ···················· | |
| B | 6577 hCoV-19/England/CAMB-73721/2020\|EPI_ISL_440285\|202 | ···················· | ···················· | ···················· | ···················· | ···················· | ···················· | |
| B | 6578 hCoV-19/England/BRIS-121AFB/2020\|EPI_ISL_440286\|20 | ···················· | ···················· | ···················· | ···················· | ···················· | ···················· | |
| B | 6579 hCoV-19/England/CAMB-73211/2020\|EPI_ISL_440287\|202 | ···················· | ···················· | ···················· | ···················· | ···················· | ···················· | |
| B | 6580 hCoV-19/England/CAMB-736F7/2020\|EPI_ISL_440288\|202 | ···················· | ···················· | ···················· | ···················· | ···················· | ···················· | |
| B | 6581 hCoV-19/England/CAMB-735FA/2020\|EPI_ISL_440289\|202 | ···················· | ···················· | ···················· | ···················· | ···················· | ···················· | |
| B | 6582 hCoV-19/England/CAMB-73475/2020\|EPI_ISL_440290\|202 | ···················· | ···················· | ···················· | ···················· | ···················· | ···················· | |
| B | 6583 hCoV-19/England/BRIS-121C40/2020\|EPI_ISL_440291\|20 | ···················· | ···················· | ···················· | ···················· | ···················· | ···················· | |
| B | 6584 hCoV-19/England/CAMB-72FFD/2020\|EPI_ISL_440293\|202 | ···················· | ···················· | ···················· | ···················· | ···················· | ···················· | |
| B | 6585 hCoV-19/England/CAMB-72F48/2020\|EPI_ISL_440295\|202 | ···················· | ···················· | ···················· | ···················· | ···················· | ···················· | |
| B | 6586 hCoV-19/England/CAMB-733F0/2020\|EPI_ISL_440296\|202 | ···················· | ···················· | ···················· | ···················· | ···················· | ···················· | |
| B | 6587 hCoV-19/England/CAMB-73141/2020\|EPI_ISL_440297\|202 | ···················· | ···················· | ···················· | ···················· | ···················· | ···················· | |
| B | 6588 hCoV-19/England/BRIS-121A19/2020\|EPI_ISL_440298\|20 | ···················· | ···················· | ···················· | ···················· | ···················· | ···················· | |
| B | 6589 hCoV-19/England/BRIS-121B25/2020\|EPI_ISL_440299\|20 | ···················· | ···················· | ···················· | ···················· | ···················· | ···················· | |
| B | 6590 hCoV-19/England/CAMB-7B8CC/2020\|EPI_ISL_441586\|202 | ···················· | ···················· | ···················· | ···················· | ···················· | ···················· | |
| B | 6591 hCoV-19/England/CAMB-7D596/2020\|EPI_ISL_441587\|202 | ···················· | ···················· | ···················· | ···················· | ···················· | ···················· | |
| B | 6592 hCoV-19/England/CAMB-7C7DD/2020\|EPI_ISL_441588\|202 | ···················· | ···················· | ···················· | ···················· | ···················· | ···················· | |
| B | 6593 hCoV-19/England/CAMB-7D2EA/2020\|EPI_ISL_441590\|202 | ···················· | ···················· | ··············T····· | ···················· | ···················· | ···················· | |
| B | 6594 hCoV-19/England/CAMB-7B7ED/2020\|EPI_ISL_441591\|202 | ···················· | ···················· | ···················· | ···················· | ···················· | ···················· | |
| B | 6595 hCoV-19/England/CAMB-7BB1E/2020\|EPI_ISL_441593\|202 | ···················· | ···················· | ···················· | ···················· | ···················· | ···················· | |
| B | 6596 hCoV-19/England/CAMB-7C861/2020\|EPI_ISL_441594\|202 | ···················· | ···················· | ···················· | ···················· | ···················· | ···················· | |
| B | 6597 hCoV-19/England/CAMB-7B817/2020\|EPI_ISL_441595\|202 | ···················· | ···················· | ···················· | ···················· | ···················· | ···················· | |
| B | 6598 hCoV-19/England/CAMB-7B7A1/2020\|EPI_ISL_441596\|202 | ···················· | ···················· | ···················· | ···················· | ···················· | ···················· | |
| B | 6599 hCoV-19/England/CAMB-7CE7E/2020\|EPI_ISL_441597\|202 | ···················· | ···················· | ···················· | ···················· | ···················· | ···················· | |
| B | 6600 hCoV-19/England/CAMB-7B7B0/2020\|EPI_ISL_441599\|202 | ···················· | ···················· | ···················· | ···················· | ···················· | ···················· | |
| B | 6601 hCoV-19/England/CAMB-809FE/2020\|EPI_ISL_442851\|202 | ···················· | ···················· | ···················· | ···················· | ···················· | ···················· | |
| B | 6602 hCoV-19/England/CAMB-7F4F1/2020\|EPI_ISL_442852\|202 | ···················· | ···················· | ···················· | ···················· | ···················· | ···················· | |
| B | 6603 hCoV-19/England/CAMB-815F9/2020\|EPI_ISL_442853\|202 | ···················· | ···················· | ···················· | ···················· | ···················· | ···················· | |
| B | 6604 hCoV-19/England/CAMB-7EF97/2020\|EPI_ISL_442854\|202 | ···················· | ···················· | ···················· | ···················· | ···················· | ···················· | |
| B | 6605 hCoV-19/England/CAMB-7EC63/2020\|EPI_ISL_442856\|202 | ···················· | ···················· | ···················· | ···················· | ···················· | ···················· | |
| B | 6606 hCoV-19/England/CAMB-81368/2020\|EPI_ISL_442857\|202 | ···················· | ···················· | ···················· | ···················· | ···················· | ···················· | |
| B | 6607 hCoV-19/England/CAMB-7F2CA/2020\|EPI_ISL_442858\|202 | ···················· | ···················· | ···················· | ···················· | ···················· | ···················· | |
| B | 6608 hCoV-19/England/CAMB-7F38B/2020\|EPI_ISL_442859\|202 | ···················· | ···················· | ···················· | ···················· | ···················· | ···················· | |
| B | 6609 hCoV-19/England/CAMB-7EDAC/2020\|EPI_ISL_442860\|202 | ···················· | ···················· | ···················· | ···················· | ···················· | ···················· | |
| B | 6610 hCoV-19/England/CAMB-812C5/2020\|EPI_ISL_442862\|202 | ···················· | ···················· | ···················· | ···················· | ···················· | ···················· | |
| B | 6611 hCoV-19/England/CAMB-7F41F/2020\|EPI_ISL_442864\|202 | ···················· | ···················· | ···················· | ···················· | ···················· | ···················· | |
| B | 6612 hCoV-19/England/CAMB-80B61/2020\|EPI_ISL_442865\|202 | ···················· | ···················· | ···················· | ···················· | ···················· | ···················· | |

| | | | | | | | | |
|---|---|---|---|---|---|---|---|---|
| B | 6613 | hCoV-19/England/CAMB-7F0DF/2020\|EPI_ISL_442870\|202 | ···················· | ···················· | ···················· | ···················· | ···················· | ···················· |
| B | 6614 | hCoV-19/England/CAMB-7EE30/2020\|EPI_ISL_442871\|202 | ···················· | ···················· | ···················· | ···················· | ···················· | ···················· |
| B | 6615 | hCoV-19/England/CAMB-7EBEE/2020\|EPI_ISL_442872\|202 | ···················· | ···················· | ···················· | ···················· | ···················· | ···················· |
| B | 6616 | hCoV-19/Wales/PHWC-2F0ED/2020\|EPI_ISL_446200\|2020- | ···················· | ···················· | ···················· | ···················· | ···················· | ···················· |
| B | 6617 | hCoV-19/Wales/PHWC-2F117/2020\|EPI_ISL_446203\|2020- | ···················· | ···················· | ···················· | ···················· | ···················· | ···················· |
| B | 6618 | hCoV-19/Wales/PHWC-2F144/2020\|EPI_ISL_446206\|2020- | ···················· | ···················· | ···················· | ···················· | ···················· | ···················· |
| B | 6619 | hCoV-19/Wales/PHWC-2F153/2020\|EPI_ISL_446207\|2020- | ···················· | ···················· | ···················· | ···················· | ···················· | ···················· |
| B | 6620 | hCoV-19/Wales/PHWC-2F180/2020\|EPI_ISL_446210\|2020- | ···················· | ···················· | ···················· | ···················· | ···················· | ···················· |
| B | 6621 | hCoV-19/Wales/PHWC-2F19F/2020\|EPI_ISL_446211\|2020- | ···················· | ···················· | ···················· | ···················· | ···················· | ···················· |
| B | 6622 | hCoV-19/Wales/PHWC-2F1AE/2020\|EPI_ISL_446212\|2020- | ···················· | ···················· | ···················· | ···················· | ···················· | ···················· |
| B | 6623 | hCoV-19/Wales/PHWC-2F1BD/2020\|EPI_ISL_446213\|2020- | ···················· | ···················· | ···················· | ···················· | ···················· | ···················· |
| B | 6624 | hCoV-19/Wales/PHWC-2F1CC/2020\|EPI_ISL_446214\|2020- | ···················· | ···················· | ···················· | ···················· | ···················· | ···················· |
| B | 6625 | hCoV-19/Wales/PHWC-2F1EA/2020\|EPI_ISL_446216\|2020- | ···················· | ···················· | ···················· | ···················· | ···················· | ···················· |
| B | 6626 | hCoV-19/Wales/PHWC-2F1F9/2020\|EPI_ISL_446217\|2020- | ···················· | ···················· | ···················· | ···················· | ···················· | ···················· |
| B | 6627 | hCoV-19/Wales/PHWC-2F214/2020\|EPI_ISL_446219\|2020- | ···················· | ···················· | ···················· | ···················· | ···················· | ···················· |
| B | 6628 | hCoV-19/Wales/PHWC-2F2D8/2020\|EPI_ISL_446230\|2020- | ···················· | ····A··············· | ···················· | ···················· | ···················· | ···················· |
| B | 6629 | hCoV-19/Wales/PHWC-2F2F6/2020\|EPI_ISL_446232\|2020- | ···················· | ···················· | ···················· | ···················· | ···················· | ···················· |
| B | 6630 | hCoV-19/Wales/PHWC-2F311/2020\|EPI_ISL_446234\|2020- | ···················· | ···················· | ···················· | ···················· | ···················· | ···················· |
| B | 6631 | hCoV-19/Wales/PHWC-2F34E/2020\|EPI_ISL_446236\|2020- | ···················· | ···················· | ···················· | ···················· | ···················· | ···················· |
| B | 6632 | hCoV-19/Wales/PHWC-2F35D/2020\|EPI_ISL_446237\|2020- | ···················· | ·············A······ | ···················· | ···················· | ···················· | ···················· |
| B | 6633 | hCoV-19/France/IDF-10014BC/2020\|EPI_ISL_447700\|202 | ···················· | ···················· | ···················· | ···················· | ···················· | ···················· |
| B | 6634 | hCoV-19/France/IDF-10080AA/2020\|EPI_ISL_447701\|202 | ···················· | ···················· | ···················· | ···················· | ···················· | ···················· |
| B | 6635 | hCoV-19/France/IDF-10019LM/2020\|EPI_ISL_447702\|202 | ···················· | ···················· | ···················· | ···················· | ···················· | ···················· |
| B | 6636 | hCoV-19/France/IDF-10018HJ/2020\|EPI_ISL_447703\|202 | ···················· | ···················· | ···················· | ···················· | ···················· | ···················· |
| B | 6637 | hCoV-19/France/IDF-10020MM/2020\|EPI_ISL_447704\|202 | ···················· | ···················· | ···················· | ···················· | ···················· | ···················· |
| B | 6638 | hCoV-19/France/IDF-50005JM/2020\|EPI_ISL_447705\|202 | ···················· | ···················· | ···················· | ···················· | ···················· | ···················· |
| B | 6639 | hCoV-19/France/IDF-50004CS/2020\|EPI_ISL_447706\|202 | ···················· | ···················· | ···················· | ···················· | ···················· | ···················· |
| B | 6640 | hCoV-19/France/IDF-10016DC/2020\|EPI_ISL_447707\|202 | ···················· | ···················· | ···················· | ···················· | ···················· | ···················· |
| B | 6641 | hCoV-19/France/IDF-10015BY/2020\|EPI_ISL_447708\|202 | ···················· | ···················· | ···················· | ···················· | ···················· | ···················· |
| B | 6642 | hCoV-19/France/IDF-10012BM/2020\|EPI_ISL_447709\|202 | ···················· | ···················· | ···················· | ···················· | ···················· | ···················· |
| B | 6643 | hCoV-19/France/IDF-10017GS/2020\|EPI_ISL_447710\|202 | ···················· | ···················· | ···················· | ···················· | ···················· | ···················· |
| B | 6644 | hCoV-19/France/IDF-10013BA/2020\|EPI_ISL_447711\|202 | ···················· | ···················· | ···················· | ···················· | ···················· | ···················· |
| B | 6645 | hCoV-19/France/IDF-10021SB/2020\|EPI_ISL_447712\|202 | ···················· | ···················· | ···················· | ···················· | ···················· | ···················· |
| B | 6646 | hCoV-19/France/IDF-10011AS/2020\|EPI_ISL_447713\|202 | ···················· | ···················· | ···················· | ···················· | ···················· | ···················· |
| B | 6647 | hCoV-19/France/IDF-10023FD/2020\|EPI_ISL_447714\|202 | ···················· | ···················· | ···················· | ···················· | ···················· | ···················· |
| B | 6648 | hCoV-19/France/IDF-10022AR/2020\|EPI_ISL_447715\|202 | ···················· | ···················· | ···················· | ···················· | ···················· | ···················· |
| B | 6649 | hCoV-19/France/IDF-50011LM/2020\|EPI_ISL_447716\|202 | ···················· | ···················· | ···················· | ···················· | ···················· | ···················· |
| B | 6650 | hCoV-19/France/IDF-10001DM/2020\|EPI_ISL_447717\|202 | ···················· | ···················· | ···················· | ···················· | ···················· | ···················· |
| B | 6651 | hCoV-19/France/IDF-50007PI/2020\|EPI_ISL_447718\|202 | ···················· | ···················· | ···················· | ···················· | ···················· | ···················· |
| B | 6652 | hCoV-19/France/IDF-10003SN/2020\|EPI_ISL_447719\|202 | ···················· | ···················· | ···················· | ···················· | ···················· | ···················· |
| B | 6653 | hCoV-19/France/IDF-10025GM/2020\|EPI_ISL_447720\|202 | ···················· | ···················· | ···················· | ···················· | ···················· | ···················· |
| B | 6654 | hCoV-19/France/IDF-10002PM/2020\|EPI_ISL_447721\|202 | ···················· | ···················· | ···················· | ···················· | ···················· | ···················· |
| B | 6655 | hCoV-19/France/IDF-10026SL/2020\|EPI_ISL_447722\|202 | ···················· | ···················· | ···················· | ···················· | ···················· | ···················· |
| B | 6656 | hCoV-19/France/IDF-30001MB/2020\|EPI_ISL_447723\|202 | ···················· | ···················· | ···················· | ···················· | ···················· | ···················· |
| B | 6657 | hCoV-19/Argentina/PAIS-A0008/2020\|EPI_ISL_430800\|2 | ···················· | ···················· | ···················· | ···················· | ···················· | ···················· |
| B | 6658 | hCoV-19/Argentina/PAIS-A0009/2020\|EPI_ISL_430801\|2 | ···················· | ···················· | ···················· | ···················· | ···················· | ···················· |
| B | 6659 | hCoV-19/Argentina/PAIS-A0011/2020\|EPI_ISL_430802\|2 | ···················· | ···················· | ···················· | ···················· | ···················· | ···················· |
| B | 6660 | hCoV-19/Argentina/PAIS-A0012/2020\|EPI_ISL_430803\|2 | ···················· | ···················· | ···················· | ···················· | ···················· | ···················· |
| B | 6661 | hCoV-19/Argentina/PAIS-A0013/2020\|EPI_ISL_430804\|2 | ···················· | ···················· | ···················· | ···················· | ···················· | ···················· |
| B | 6662 | hCoV-19/Argentina/PAIS-A0014/2020\|EPI_ISL_430805\|2 | ···················· | ···················· | ···················· | ···················· | ···················· | ···················· |
| B | 6663 | hCoV-19/Argentina/PAIS-A0016/2020\|EPI_ISL_430807\|2 | ···················· | ···················· | ···················· | ···················· | ···················· | ···················· |
| B | 6664 | hCoV-19/Argentina/PAIS-A0017/2020\|EPI_ISL_430808\|2 | ···················· | ···················· | ···················· | ···················· | ···················· | ···················· |
| B | 6665 | hCoV-19/Argentina/PAIS-A0018/2020\|EPI_ISL_430809\|2 | ···················· | ···················· | ···················· | ···················· | ···················· | ···················· |
| B | 6666 | hCoV-19/Thailand/TNIC-1889/2020\|EPI_ISL_430837\|202 | ···················· | ···················· | ···················· | ···················· | ···················· | ···················· |
| B | 6667 | hCoV-19/Sweden/20-08150/2020\|EPI_ISL_430850\|2020-0 | ···················· | ···················· | ···················· | ···················· | ···················· | ···················· |
| B | 6668 | hCoV-19/Sweden/20-08152/2020\|EPI_ISL_430851\|2020-0 | ···················· | ···················· | ···················· | ···················· | ···················· | ···················· |
| B | 6669 | hCoV-19/Sweden/20-08154/2020\|EPI_ISL_430852\|2020-0 | ···················· | ···················· | ···················· | ···················· | ···················· | ···················· |
| B | 6670 | hCoV-19/Sweden/20-08167/2020\|EPI_ISL_430853\|2020-0 | ···················· | ···················· | ···················· | ···················· | ···················· | ···················· |
| B | 6671 | hCoV-19/Sweden/20-08171/2020\|EPI_ISL_430854\|2020-0 | ···················· | ···················· | ···················· | ···················· | ···················· | ···················· |
| B | 6672 | hCoV-19/Sweden/20-08172/2020\|EPI_ISL_430855\|2020-0 | ···················· | ···················· | ···················· | ···················· | ···················· | ···················· |
| B | 6673 | hCoV-19/Sweden/20-08687/2020\|EPI_ISL_430856\|2020-0 | ···················· | ····A··············· | ···················· | ···················· | ···················· | ···················· |
| B | 6674 | hCoV-19/Sweden/20-08691/2020\|EPI_ISL_430857\|2020-0 | ···················· | ···················· | ···················· | ···················· | ···················· | ···················· |
| B | 6675 | hCoV-19/Sweden/20-08714/2020\|EPI_ISL_430858\|2020-0 | ···················· | ···················· | ···················· | ···················· | ···················· | ···················· |
| B | 6676 | hCoV-19/Sweden/20-08717/2020\|EPI_ISL_430859\|2020-0 | ···················· | ···················· | ···················· | ···················· | ···················· | ···················· |
| B | 6677 | hCoV-19/Sweden/20-50043/2020\|EPI_ISL_430860\|2020-0 | ···················· | ···················· | ···················· | ···················· | ···················· | ···················· |
| B | 6678 | hCoV-19/Sweden/20-50129/2020\|EPI_ISL_430861\|2020-0 | ···················· | ···················· | ···················· | ···················· | ···················· | ···················· |
| B | 6679 | hCoV-19/Sweden/20-50185/2020\|EPI_ISL_430862\|2020-0 | ···················· | ···················· | ···················· | ···················· | ···················· | ···················· |
| B | 6680 | hCoV-19/Sweden/20-50231/2020\|EPI_ISL_430863\|2020-0 | ···················· | ···················· | ···················· | ···················· | ···················· | ···················· |
| B | 6681 | hCoV-19/Sweden/20-50360/2020\|EPI_ISL_430864\|2020-0 | ···················· | ···················· | ···················· | ···················· | ···················· | ···················· |
| B | 6682 | hCoV-19/USA/WA-UW-517/2020\|EPI_ISL_430867\|2020-03- | ···················· | ···················· | ···················· | ···················· | ···················· | ···················· |
| B | 6683 | hCoV-19/USA/WA-UW-5177/2020\|EPI_ISL_430868\|2020-04 | ···················· | ···················· | ···················· | ···················· | ···················· | ···················· |
| B | 6684 | hCoV-19/USA/WA-UW-5178/2020\|EPI_ISL_430869\|2020-04 | ···················· | ···················· | ···················· | ···················· | ···················· | ···················· |
| B | 6685 | hCoV-19/USA/WA-UW-518/2020\|EPI_ISL_430870\|2020-03- | ···················· | ···················· | ···················· | ···················· | ···················· | ···················· |
| B | 6686 | hCoV-19/USA/WA-UW-5181/2020\|EPI_ISL_430871\|2020-04 | ···················· | ···················· | ···················· | ···················· | ···················· | ···················· |
| B | 6687 | hCoV-19/USA/WA-UW-602/2020\|EPI_ISL_430894\|2020-03- | ···················· | ···················· | ···················· | ···················· | ···················· | ···················· |
| B | 6688 | hCoV-19/USA/WA-UW-604/2020\|EPI_ISL_430895\|2020-03- | ···················· | ···················· | ···················· | ···················· | ···················· | ···················· |
| B | 6689 | hCoV-19/USA/UN-UW-5199/2020\|EPI_ISL_430897\|2020-04 | ···················· | ···················· | ···················· | ···················· | ···················· | ···················· |
| B | 6690 | hCoV-19/USA/WA-UW-5200/2020\|EPI_ISL_430898\|2020-04 | ···················· | ···················· | ···················· | ···················· | ···················· | ···················· |

| | | | | | | | | |
|---|---|---|---|---|---|---|---|---|
| B | 6691 hCoV-19/USA/WA-UW-5203/2020\|EPI_ISL_430899\|2020-04 | ···················· | ···················· | ···················· | ···················· | ···················· | ···················· | |
| B | 6692 hCoV-19/England/NOTT-10F98D/2020\|EPI_ISL_433066\|20 | ···················· | ···················· | ···················· | ···················· | ···················· | ···················· | |
| B | 6693 hCoV-19/Scotland/EDB1518/2020\|EPI_ISL_433068\|2020- | ···················· | ···················· | ···················· | ···················· | ···················· | ···················· | |
| B | 6694 hCoV-19/Scotland/EDB1519/2020\|EPI_ISL_433069\|2020- | ···················· | ···················· | ···················· | ···················· | ···················· | ···················· | |
| B | 6695 hCoV-19/Scotland/EDB1523/2020\|EPI_ISL_433070\|2020- | ···················· | ·A·················· | ···················· | ···················· | ···················· | ···················· | |
| B | 6696 hCoV-19/Scotland/EDB1525/2020\|EPI_ISL_433071\|2020- | ···················· | ···················· | ···················· | ···················· | ···················· | ···················· | |
| B | 6697 hCoV-19/Scotland/EDB1526/2020\|EPI_ISL_433072\|2020- | ···················· | ·A·················· | ···················· | ···················· | ···················· | ···················· | |
| B | 6698 hCoV-19/Scotland/EDB1529/2020\|EPI_ISL_433073\|2020- | ···················· | ···················· | ···················· | ···················· | ···················· | ···················· | |
| B | 6699 hCoV-19/Scotland/EDB153/2020\|EPI_ISL_433074\|2020-0 | ···················· | ···················· | ···················· | ···················· | ···················· | ···················· | |
| B | 6700 hCoV-19/Scotland/EDB1536/2020\|EPI_ISL_433075\|2020- | ···················· | ·A·················· | ···················· | ···················· | ···················· | ···················· | |
| B | 6701 hCoV-19/Scotland/EDB154/2020\|EPI_ISL_433076\|2020-0 | ···················· | ···················· | ···················· | ···················· | ···················· | ···················· | |
| B | 6702 hCoV-19/Scotland/EDB1541/2020\|EPI_ISL_433077\|2020- | ···················· | ·A·················· | ···················· | ···················· | ···················· | ···················· | |
| B | 6703 hCoV-19/Scotland/EDB1544/2020\|EPI_ISL_433078\|2020- | ···················· | ·A·················· | ···················· | ···················· | ···················· | ···················· | |
| B | 6704 hCoV-19/Scotland/EDB1550/2020\|EPI_ISL_433079\|2020- | ···················· | ···················· | ···················· | ···················· | ···················· | ···················· | |
| B | 6705 hCoV-19/Scotland/EDB1554/2020\|EPI_ISL_433080\|2020- | ···················· | ·A·················· | ···················· | ···················· | ···················· | ···················· | |
| B | 6706 hCoV-19/Scotland/EDB1559/2020\|EPI_ISL_433081\|2020- | ···················· | ·A·················· | ···················· | ···················· | ···················· | ···················· | |
| B | 6707 hCoV-19/Scotland/EDB1565/2020\|EPI_ISL_433082\|2020- | ···················· | ···················· | ···················· | ···················· | ···················· | ···················· | |
| B | 6708 hCoV-19/Scotland/EDB1566/2020\|EPI_ISL_433083\|2020- | ···················· | ···················· | ···················· | ···················· | ···················· | ···················· | |
| B | 6709 hCoV-19/Scotland/EDB1568/2020\|EPI_ISL_433084\|2020- | ···················· | ···················· | ···················· | ···················· | ···················· | ···················· | |
| B | 6710 hCoV-19/Scotland/EDB1580/2020\|EPI_ISL_433085\|2020- | ···················· | ·A·················· | ···················· | ···················· | ···················· | ···················· | |
| B | 6711 hCoV-19/Scotland/EDB1581/2020\|EPI_ISL_433086\|2020- | ···················· | ···················· | ···················· | ···················· | ···················· | ···················· | |
| B | 6712 hCoV-19/Scotland/EDB1582/2020\|EPI_ISL_433087\|2020- | ···················· | ···················· | ···················· | ···················· | ···················· | ···················· | |
| B | 6713 hCoV-19/Scotland/EDB1583/2020\|EPI_ISL_433088\|2020- | ···················· | ···················· | ···················· | ···················· | ···················· | ···················· | |
| B | 6720 hCoV-19/Scotland/EDB2403/2020\|EPI_ISL_439200\|2020- | ···················· | ···················· | ···················· | ···················· | ···················· | ···················· | |
| B | 6721 hCoV-19/Scotland/EDB2405/2020\|EPI_ISL_439201\|2020- | ···················· | ···················· | ···················· | ···················· | ···················· | ···················· | |
| B | 6722 hCoV-19/Scotland/EDB2409/2020\|EPI_ISL_439202\|2020- | ···················· | ···················· | ···················· | ···················· | ···················· | ···················· | |
| B | 6723 hCoV-19/Scotland/EDB2412/2020\|EPI_ISL_439203\|2020- | ···················· | ···················· | ···················· | ···················· | ···················· | ···················· | |
| B | 6724 hCoV-19/Scotland/EDB2416/2020\|EPI_ISL_439204\|2020- | ···················· | ···················· | ···················· | ···················· | ···················· | ···················· | |
| B | 6725 hCoV-19/Scotland/EDB2419/2020\|EPI_ISL_439205\|2020- | ···················· | ···················· | ···················· | ···················· | ···················· | ···················· | |
| B | 6726 hCoV-19/Scotland/EDB2432/2020\|EPI_ISL_439206\|2020- | ···················· | ···················· | ···················· | ···················· | ···················· | ···················· | |
| B | 6727 hCoV-19/Scotland/EDB2435/2020\|EPI_ISL_439207\|2020- | ···················· | ·A·················· | ···················· | ···················· | ···················· | ···················· | |
| B | 6728 hCoV-19/Scotland/EDB2440/2020\|EPI_ISL_439208\|2020- | ···················· | ···················· | ···················· | ···················· | ···················· | ···················· | |
| B | 6729 hCoV-19/Scotland/EDB2445/2020\|EPI_ISL_439209\|2020- | ···················· | ···················· | ···················· | ···················· | ···················· | ···················· | |
| B | 6730 hCoV-19/Scotland/EDB2476/2020\|EPI_ISL_439220\|2020- | ···················· | ···················· | ···················· | ···················· | ···················· | ···················· | |
| B | 6731 hCoV-19/Scotland/EDB2483/2020\|EPI_ISL_439221\|2020- | ·T·················· | ···················· | ···················· | ···················· | ···················· | ···················· | |
| B | 6732 hCoV-19/Scotland/EDB2561/2020\|EPI_ISL_439230\|2020- | ···················· | ···················· | ···················· | ···················· | ···················· | ···················· | |
| B | 6733 hCoV-19/Scotland/EDB2563/2020\|EPI_ISL_439231\|2020- | ···················· | ···················· | ···················· | ···················· | ···················· | ···················· | |
| B | 6734 hCoV-19/Scotland/EDB2567/2020\|EPI_ISL_439232\|2020- | ···················· | ···················· | ···················· | ···················· | ···················· | ···················· | |
| B | 6735 hCoV-19/Scotland/EDB2579/2020\|EPI_ISL_439233\|2020- | ···················· | ···················· | ···················· | ···················· | ···················· | ···················· | |
| B | 6736 hCoV-19/Scotland/EDB2592/2020\|EPI_ISL_439234\|2020- | ···················· | ·A·················· | ···················· | ···················· | ···················· | ···················· | |
| B | 6737 hCoV-19/Scotland/EDB2594/2020\|EPI_ISL_439235\|2020- | ···················· | ·A·················· | ···················· | ···················· | ···················· | ···················· | |
| B | 6738 hCoV-19/Scotland/EDB2597/2020\|EPI_ISL_439236\|2020- | ···················· | ···················· | ···················· | ···················· | ···················· | ···················· | |
| B | 6739 hCoV-19/Scotland/EDB2599/2020\|EPI_ISL_439237\|2020- | ···················· | ···················· | ···················· | ···················· | ···················· | ···················· | |
| B | 6740 hCoV-19/Scotland/EDB2605/2020\|EPI_ISL_439238\|2020- | ···················· | ·A·················· | ···················· | ···················· | ···················· | ···················· | |
| B | 6741 hCoV-19/Scotland/EDB2606/2020\|EPI_ISL_439239\|2020- | ···················· | ···················· | ···················· | ···················· | ···················· | ···················· | |
| B | 6742 hCoV-19/Scotland/EDB2607/2020\|EPI_ISL_439240\|2020- | ···················· | ···················· | ···················· | ···················· | ···················· | ···················· | |
| B | 6743 hCoV-19/Scotland/EDB2608/2020\|EPI_ISL_439241\|2020- | ···················· | ·A·················· | ···················· | ···················· | ···················· | ···················· | |
| B | 6744 hCoV-19/Scotland/EDB2609/2020\|EPI_ISL_439242\|2020- | ···················· | ···················· | ···················· | ···················· | ···················· | ···················· | |
| B | 6745 hCoV-19/Scotland/EDB2614/2020\|EPI_ISL_439243\|2020- | ···················· | ···················· | ···················· | ···················· | ···················· | ···················· | |
| B | 6746 hCoV-19/Scotland/EDB2620/2020\|EPI_ISL_439244\|2020- | ···················· | ···················· | ···················· | ···················· | ···················· | ···················· | |
| B | 6747 hCoV-19/Scotland/EDB2627/2020\|EPI_ISL_439245\|2020- | ···················· | ···················· | ···················· | ···················· | ···················· | ···················· | |
| B | 6748 hCoV-19/Scotland/EDB2631/2020\|EPI_ISL_439246\|2020- | ···················· | ···················· | ···················· | ···················· | ···················· | ···················· | |
| B | 6749 hCoV-19/Scotland/EDB2635/2020\|EPI_ISL_439247\|2020- | ···················· | ···················· | ···················· | ···················· | ···················· | ···················· | |
| B | 6750 hCoV-19/Scotland/EDB2638/2020\|EPI_ISL_439248\|2020- | ···················· | ·A·················· | ···················· | ···················· | ···················· | ···················· | |
| B | 6751 hCoV-19/Scotland/EDB2650/2020\|EPI_ISL_439249\|2020- | ···················· | ···················· | ···················· | ···················· | ···················· | ···················· | |
| B | 6752 hCoV-19/Scotland/EDB2656/2020\|EPI_ISL_439250\|2020- | ···················· | ···················· | ···················· | ···················· | ···················· | ···················· | |
| B | 6753 hCoV-19/Scotland/EDB2659/2020\|EPI_ISL_439251\|2020- | ···················· | ·A·················· | ···················· | ···················· | ···················· | ···················· | |
| B | 6754 hCoV-19/Scotland/EDB2661/2020\|EPI_ISL_439253\|2020- | ···················· | ···················· | ···················· | ···················· | ···················· | ···················· | |
| B | 6755 hCoV-19/Scotland/EDB2662/2020\|EPI_ISL_439254\|2020- | ···················· | ·A·················· | ···················· | ···················· | ···················· | ···················· | |
| B | 6756 hCoV-19/Scotland/EDB2663/2020\|EPI_ISL_439255\|2020- | ···················· | ·A·················· | ···················· | ···················· | ···················· | ···················· | |
| B | 6757 hCoV-19/Scotland/EDB2666/2020\|EPI_ISL_439256\|2020- | ···················· | ···················· | ···················· | ···················· | ···················· | ···················· | |
| B | 6758 hCoV-19/Scotland/EDB2670/2020\|EPI_ISL_439257\|2020- | ···················· | ···················· | ···················· | ···················· | ···················· | ···················· | |
| B | 6759 hCoV-19/Scotland/EDB2673/2020\|EPI_ISL_439258\|2020- | ···················· | ·A·················· | ···················· | ···················· | ···················· | ···················· | |
| B | 6760 hCoV-19/Scotland/EDB2674/2020\|EPI_ISL_439259\|2020- | ···················· | ···················· | ···················· | ···················· | ···················· | ···················· | |
| B | 6761 hCoV-19/Scotland/EDB2678/2020\|EPI_ISL_439261\|2020- | ···················· | ·A·················· | ···················· | ···················· | ···················· | ··A················· | |
| B | 6762 hCoV-19/Scotland/EDB2679/2020\|EPI_ISL_439262\|2020- | ···················· | ···················· | ···················· | ···················· | ···················· | ···················· | |
| B | 6763 hCoV-19/Scotland/EDB2683/2020\|EPI_ISL_439263\|2020- | ···················· | ···················· | ···················· | ···················· | ···················· | ···················· | |
| B | 6764 hCoV-19/Scotland/EDB2684/2020\|EPI_ISL_439264\|2020- | ···················· | ···················· | ···················· | ···················· | ···················· | ···················· | |
| B | 6766 hCoV-19/Scotland/EDB3454/2020\|EPI_ISL_439266\|2020- | ···················· | ···················· | ···················· | ···················· | ···················· | ···················· | |
| B | 6767 hCoV-19/Scotland/EDB3456/2020\|EPI_ISL_439267\|2020- | ···················· | ···················A | ···················· | ···················· | ···················· | ···················· | 1 |
| B | 6768 hCoV-19/Scotland/EDB3495/2020\|EPI_ISL_439268\|2020- | ···················· | ···················A | ···················· | ···················· | ···················· | ···················· | 1 |
| B | 6769 hCoV-19/Scotland/EDB3496/2020\|EPI_ISL_439269\|2020- | ···················· | ···················A | ···················· | ···················· | ···················· | ···················· | 1 |
| B | 6770 hCoV-19/England/CAMB-734A2/2020\|EPI_ISL_440202\|202 | ···················· | ···················· | ···················· | ···················· | ···················· | ···················· | |
| B | 6771 hCoV-19/England/CAMB-731AB/2020\|EPI_ISL_440203\|202 | ···················· | ···················· | ···················· | ···················· | ···················· | ···················· | |
| B | 6772 hCoV-19/England/BRIS-121BDA/2020\|EPI_ISL_440206\|20 | ···················· | ···················· | ···················· | ···················· | ···················· | ···················· | |
| B | 6773 hCoV-19/England/CAMB-7316F/2020\|EPI_ISL_440207\|202 | ···················· | ···················· | ···················· | ···················· | ···················· | ···················· | |
| B | 6774 hCoV-19/England/CAMB-73624/2020\|EPI_ISL_440213\|202 | ···················· | ···················· | ···················· | ···················· | ···················· | ···················· | |
| B | 6775 hCoV-19/England/BRIS-121800/2020\|EPI_ISL_440214\|20 | ···················· | ···················· | ···················· | ···················· | ···················· | ···················· | |

| | | | | | | | | |
|---|---|---|---|---|---|---|---|---|
| B | 6776 | hCoV-19/England/CAMB-7323F/2020\|EPI_ISL_440217\|202 | .................... | .................... | .................... | .................... | .................... | .................... |
| B | 6777 | hCoV-19/England/BRIS-121C9B/2020\|EPI_ISL_440218\|20 | .................... | .................... | .................... | .................... | .................... | .................... |
| B | 6778 | hCoV-19/England/BRIS-12184C/2020\|EPI_ISL_440219\|20 | .................... | .................... | .................... | .................... | .................... | .................... |
| B | 6779 | hCoV-19/England/BRIS-121AEC/2020\|EPI_ISL_440222\|20 | .................... | .................... | .................... | .................... | .................... | .................... |
| B | 6780 | hCoV-19/England/CAMB-7342A/2020\|EPI_ISL_440223\|202 | .................... | .................... | .................... | .................... | .................... | .................... |
| B | 6781 | hCoV-19/England/BRIS-12176D/2020\|EPI_ISL_440224\|20 | .................... | .................... | .................... | .................... | .................... | .................... |
| B | 6782 | hCoV-19/England/BRIS-12175E/2020\|EPI_ISL_440225\|20 | .................... | .................... | .................... | .................... | .................... | .................... |
| B | 6783 | hCoV-19/England/CAMB-73008/2020\|EPI_ISL_440226\|202 | .................... | .................... | .................... | .................... | .................... | .................... |
| B | 6784 | hCoV-19/England/CAMB-73484/2020\|EPI_ISL_440231\|202 | .................... | .................... | .................... | .................... | .................... | .................... |
| B | 6785 | hCoV-19/England/BRIS-121C5F/2020\|EPI_ISL_440232\|20 | .................... | .................... | .................... | .................... | .................... | .................... |
| B | 6786 | hCoV-19/England/BRIS-121976/2020\|EPI_ISL_440233\|20 | .................... | .................... | .................... | .................... | .................... | .................... |
| B | 6787 | hCoV-19/England/CAMB-7367F/2020\|EPI_ISL_440234\|202 | .................... | .................... | .................... | .................... | .................... | .................... |
| B | 6788 | hCoV-19/England/BRIS-121B16/2020\|EPI_ISL_440236\|20 | .................... | .................... | .................... | .................... | .................... | .................... |
| B | 6789 | hCoV-19/England/CAMB-73080/2020\|EPI_ISL_440237\|202 | .................... | .................... | .................... | .................... | .................... | .................... |
| B | 6790 | hCoV-19/England/CAMB-73017/2020\|EPI_ISL_440239\|202 | .................... | .................... | .................... | .................... | .................... | .................... |
| B | 6791 | hCoV-19/England/BRIS-121CD7/2020\|EPI_ISL_440241\|20 | .................... | .................... | .................... | .................... | .................... | .................... |
| B | 6792 | hCoV-19/England/BRIS-121C13/2020\|EPI_ISL_440243\|20 | .................... | .................... | .................... | .................... | .................... | .................... |
| B | 6793 | hCoV-19/England/CAMB-732E4/2020\|EPI_ISL_440244\|202 | .................... | .A.................. | .................... | .................... | .................... | .................... |
| B | 6794 | hCoV-19/England/CAMB-730F9/2020\|EPI_ISL_440246\|202 | .................... | .................... | .................... | .................... | .................... | .................... |
| B | 6795 | hCoV-19/England/BRIS-1219A3/2020\|EPI_ISL_440248\|20 | .................... | .................... | .................... | .................... | .................... | .................... |
| B | 6796 | hCoV-19/England/BRIS-12186A/2020\|EPI_ISL_440250\|20 | .................... | .................... | .................... | .................... | .................... | .................... |
| B | 6797 | hCoV-19/England/CAMB-72F39/2020\|EPI_ISL_440254\|202 | .................... | .................... | .................... | .................... | .................... | .................... |
| B | 6798 | hCoV-19/England/CAMB-7C685/2020\|EPI_ISL_441547\|202 | .................... | .................... | .................... | .................... | .................... | .................... |
| B | 6799 | hCoV-19/England/CAMB-7D0E0/2020\|EPI_ISL_441549\|202 | .................... | .................... | .................... | .................... | .................... | .................... |
| B | 6800 | hCoV-19/England/CAMB-7D165/2020\|EPI_ISL_441550\|202 | .................... | .................... | .................... | .................... | .................... | .................... |
| B | 6801 | hCoV-19/England/CAMB-7C500/2020\|EPI_ISL_441551\|202 | .................... | .................... | .................... | .................... | .................... | .................... |
| B | 6802 | hCoV-19/England/CAMB-7D6EE/2020\|EPI_ISL_441552\|202 | .................... | .................... | .................... | .................... | .................... | .................... |
| B | 6803 | hCoV-19/England/CAMB-7CCDE/2020\|EPI_ISL_441554\|202 | .................... | .................... | .................... | .................... | .................... | .................... |
| B | 6804 | hCoV-19/England/CAMB-7CA5C/2020\|EPI_ISL_441555\|202 | .................... | .................... | .................... | .................... | .................... | .................... |
| B | 6805 | hCoV-19/England/CAMB-7D2BD/2020\|EPI_ISL_441556\|202 | .................... | .................... | .................... | .................... | .................... | .................... |
| B | 6806 | hCoV-19/England/CAMB-7BAC6/2020\|EPI_ISL_441557\|202 | .................... | .................... | .................... | .................... | .................... | .................... |
| B | 6807 | hCoV-19/England/CAMB-7CC83/2020\|EPI_ISL_441558\|202 | .................... | .................... | .................... | .................... | .................... | .................... |
| B | 6808 | hCoV-19/England/CAMB-7CF11/2020\|EPI_ISL_441559\|202 | .................... | .................... | .................... | .................... | .................... | .................... |
| B | 6809 | hCoV-19/England/CAMB-7D305/2020\|EPI_ISL_441560\|202 | .................... | .................... | .................... | .................... | .................... | .................... |
| B | 6810 | hCoV-19/England/CAMB-7DBC1/2020\|EPI_ISL_441561\|202 | .................... | .................... | .................... | .................... | .................... | .................... |
| B | 6811 | hCoV-19/England/CAMB-7C667/2020\|EPI_ISL_441562\|202 | .................... | .................... | .................... | .................... | .................... | .................... |
| B | 6812 | hCoV-19/England/CAMB-7D684/2020\|EPI_ISL_441563\|202 | .................... | .................... | .................... | .................... | .................... | .................... |
| B | 6813 | hCoV-19/England/CAMB-7CEBA/2020\|EPI_ISL_441564\|202 | .................... | .................... | .................... | .................... | .................... | .................... |
| B | 6814 | hCoV-19/England/CAMB-7D3AB/2020\|EPI_ISL_441565\|202 | .................... | .................... | .................... | .................... | .................... | .................... |
| B | 6815 | hCoV-19/England/CAMB-7D833/2020\|EPI_ISL_441566\|202 | .................... | .................... | .................... | .................... | .................... | .................... |
| B | 6816 | hCoV-19/England/CAMB-7B97E/2020\|EPI_ISL_441569\|202 | .................... | .................... | .................... | .................... | .................... | .................... |
| B | 6817 | hCoV-19/England/CAMB-7C7FB/2020\|EPI_ISL_441571\|202 | .................... | .................... | .................... | .................... | .................... | .................... |
| B | 6818 | hCoV-19/England/CAMB-7E00D/2020\|EPI_ISL_441572\|202 | .................... | .................... | .................... | .................... | .................... | .................... |
| B | 6819 | hCoV-19/England/CAMB-7C676/2020\|EPI_ISL_441573\|202 | .................... | .................... | .................... | .................... | .................... | .................... |
| B | 6820 | hCoV-19/England/CAMB-7B844/2020\|EPI_ISL_441574\|202 | .................... | .................... | .................... | .................... | .................... | .................... |
| B | 6821 | hCoV-19/England/CAMB-7CD53/2020\|EPI_ISL_441575\|202 | .................... | .................... | .................... | .................... | .................... | .................... |
| B | 6822 | hCoV-19/England/CAMB-7DC46/2020\|EPI_ISL_441576\|202 | .................... | .................... | .................... | .................... | .................... | .................... |
| B | 6823 | hCoV-19/England/CAMB-7C6EF/2020\|EPI_ISL_441577\|202 | .................... | .................... | .................... | .................... | .................... | .................... |
| B | 6824 | hCoV-19/England/CAMB-7CD17/2020\|EPI_ISL_441578\|202 | .................... | .................... | .................... | .................... | .................... | .................... |
| B | 6825 | hCoV-19/England/CAMB-7C588/2020\|EPI_ISL_441579\|202 | .................... | .................... | .................... | .................... | .................... | .................... |
| B | 6826 | hCoV-19/England/CAMB-7DA4C/2020\|EPI_ISL_441582\|202 | .................... | .................... | .................... | .................... | .................... | .................... |
| B | 6827 | hCoV-19/England/CAMB-7B923/2020\|EPI_ISL_441583\|202 | .................... | .................... | .................... | .................... | .................... | .................... |
| B | 6828 | hCoV-19/England/CAMB-7C412/2020\|EPI_ISL_441584\|202 | .................... | .................... | .................... | .................... | .................... | .................... |
| B | 6829 | hCoV-19/England/CAMB-7D341/2020\|EPI_ISL_441585\|202 | .................... | .................... | .................... | .................... | .................... | .................... |
| B | 6830 | hCoV-19/England/BRIS-1235E9/2020\|EPI_ISL_442810\|20 | .................... | .................... | .................... | .................... | .................... | .................... |
| B | 6831 | hCoV-19/England/CAMB-7D04A/2020\|EPI_ISL_442811\|202 | .................... | .................... | .................... | .................... | .................... | .................... |
| B | 6832 | hCoV-19/England/CAMB-7D0A4/2020\|EPI_ISL_442812\|202 | .................... | .................... | .................... | .................... | .................... | .................... |
| B | 6833 | hCoV-19/England/CAMB-7CA01/2020\|EPI_ISL_442813\|202 | .................... | .................... | .................... | .................... | .................... | .................... |
| B | 6834 | hCoV-19/England/CAMB-7BF12/2020\|EPI_ISL_442814\|202 | .................... | .................... | .................... | .................... | .................... | .................... |
| B | 6835 | hCoV-19/England/CAMB-7C70A/2020\|EPI_ISL_442815\|202 | .................... | .................... | .................... | .................... | .................... | .................... |
| B | 6836 | hCoV-19/England/CAMB-7D217/2020\|EPI_ISL_442816\|202 | .................... | .................... | .................... | .................... | .................... | .................... |
| B | 6837 | hCoV-19/England/CAMB-7C904/2020\|EPI_ISL_442817\|202 | .................... | .................... | .................... | .................... | .................... | .................... |
| B | 6838 | hCoV-19/England/CAMB-7BB87/2020\|EPI_ISL_442818\|202 | .................... | .................... | .................... | .................... | .................... | .................... |
| B | 6839 | hCoV-19/England/CAMB-7C940/2020\|EPI_ISL_442819\|202 | .................... | .................... | .................... | .................... | .................... | .................... |
| B | 6840 | hCoV-19/England/CAMB-80888/2020\|EPI_ISL_442830\|202 | .................... | .................... | .................... | .................... | .................... | .................... |
| B | 6841 | hCoV-19/England/CAMB-7F1AF/2020\|EPI_ISL_442831\|202 | .................... | .................... | .................... | .................... | .................... | .................... |
| B | 6842 | hCoV-19/England/CAMB-80DA7/2020\|EPI_ISL_442832\|202 | .................... | .................... | .................... | .................... | .................... | .................... |
| B | 6843 | hCoV-19/England/CAMB-7F181/2020\|EPI_ISL_442833\|202 | .................... | .................... | .................... | .................... | .................... | .................... |
| B | 6844 | hCoV-19/England/CAMB-7ED15/2020\|EPI_ISL_442834\|202 | .................... | .................... | .................... | .................... | .................... | .................... |
| B | 6845 | hCoV-19/England/CAMB-7F3B8/2020\|EPI_ISL_442835\|202 | .................... | .................... | .................... | .................... | .................... | .................... |
| B | 6846 | hCoV-19/England/CAMB-7F224/2020\|EPI_ISL_442839\|202 | .................... | .................... | .................... | .................... | .................... | .................... |
| B | 6847 | hCoV-19/England/CAMB-7F260/2020\|EPI_ISL_442841\|202 | .................... | .................... | .................... | .................... | .................... | .................... |
| B | 6848 | hCoV-19/England/CAMB-7EC09/2020\|EPI_ISL_442842\|202 | .................... | .................... | .................... | .................... | .................... | .................... |
| B | 6849 | hCoV-19/England/CAMB-80E2C/2020\|EPI_ISL_442844\|202 | .................... | .................... | .................... | .................... | .................... | .................... |
| B | 6850 | hCoV-19/England/CAMB-815BD/2020\|EPI_ISL_442845\|202 | .................... | .................... | .................... | .................... | .................... | .................... |
| B | 6851 | hCoV-19/England/CAMB-8126B/2020\|EPI_ISL_442846\|202 | .................... | .................... | .................... | .................... | .................... | .................... |
| B | 6852 | hCoV-19/England/CAMB-7F039/2020\|EPI_ISL_442847\|202 | .................... | .................... | .................... | .................... | .................... | .................... |
| B | 6853 | hCoV-19/England/CAMB-80B16/2020\|EPI_ISL_442848\|202 | .................... | .................... | .................... | .................... | .................... | .................... |

```
B  6854 hCoV-19/England/CAMB-7EAE1/2020|EPI_ISL_442849|202  .................... .................... .................... .................... .................... ....................
B  6855 hCoV-19/England/CAMB-7EEC7/2020|EPI_ISL_442850|202  .................... .................... .................... .................... .................... ....................
B  6856 hCoV-19/England/CAMB-81562/2020|EPI_ISL_442863|202  .................... .................... .................... .................... .................... ....................
B  6857 hCoV-19/England/CAMB-7EBFD/2020|EPI_ISL_442866|202  .................... .................... .................... .................... .................... ....................
B  6858 hCoV-19/England/CAMB-80976/2020|EPI_ISL_442867|202  .................... .................... .................... .................... .................... ....................
B  6859 hCoV-19/England/CAMB-7F28E/2020|EPI_ISL_442868|202  .................... .................... .................... .................... .................... ....................
B  6860 hCoV-19/England/CAMB-7ED24/2020|EPI_ISL_442869|202  .................... .................... .................... .................... .................... ....................
B  6861 hCoV-19/England/CAMB-80994/2020|EPI_ISL_442873|202  .................... .................... .................... .................... .................... ....................
B  6862 hCoV-19/England/CAMB-7ED8E/2020|EPI_ISL_442874|202  .................... .................... .................... .................... .................... ....................
B  6863 hCoV-19/England/CAMB-7EE8B/2020|EPI_ISL_442875|202  .................... .................... .................... .................... .................... ....................
B  6864 hCoV-19/England/CAMB-812A7/2020|EPI_ISL_442876|202  .................... .................... .................... .................... .................... ....................
B  6865 hCoV-19/England/CAMB-80B43/2020|EPI_ISL_442878|202  .................... .................... .................... .................... .................... ....................
B  6866 hCoV-19/England/CAMB-7EDF7/2020|EPI_ISL_442879|202  .................... .................... .................... .................... .................... ....................
B  6867 hCoV-19/England/CAMB-80B9E/2020|EPI_ISL_442880|202  .................... .................... .................... .................... .................... ....................
B  6868 hCoV-19/England/CAMB-7ECDC/2020|EPI_ISL_442881|202  .................... .................... .................... .................... .................... ....................
B  6869 hCoV-19/England/CAMB-80B25/2020|EPI_ISL_442882|202  .................... .................... .................... .................... .................... ....................
B  6870 hCoV-19/England/CAMB-808E2/2020|EPI_ISL_442883|202  .................... .................... .................... .................... .................... ....................
B  6871 hCoV-19/England/CAMB-7F233/2020|EPI_ISL_442884|202  .................... .................... .................... .................... .................... ....................
B  6872 hCoV-19/England/CAMB-808C4/2020|EPI_ISL_442885|202  .................... .................... .................... .................... .................... ....................
B  6873 hCoV-19/England/CAMB-80C8C/2020|EPI_ISL_442886|202  .................... .A.................. .................... .................... .................... ....................
B  6874 hCoV-19/England/CAMB-7EC90/2020|EPI_ISL_442888|202  .................... .................... .................... .................... .................... ....................
B  6875 hCoV-19/England/CAMB-7F154/2020|EPI_ISL_442889|202  .................... .................... .................... .................... .................... ....................
B  6876 hCoV-19/England/CAMB-7F084/2020|EPI_ISL_442890|202  .................... .................... .................... .................... .................... ....................
B  6877 hCoV-19/England/CAMB-7EAF0/2020|EPI_ISL_442891|202  .................... .................... .................... .................... .................... ....................
B  6878 hCoV-19/England/CAMB-7ECAF/2020|EPI_ISL_442892|202  .................... .................... .................... .................... .................... ....................
B  6879 hCoV-19/England/CAMB-8133B/2020|EPI_ISL_442894|202  .................... .................... .................... .................... .................... ....................
B  6880 hCoV-19/England/CAMB-80897/2020|EPI_ISL_442895|202  .................... .................... .................... .................... .................... ....................
B  6881 hCoV-19/England/CAMB-7EE7C/2020|EPI_ISL_442897|202  .................... .................... .................... .................... .................... ....................
B  6882 hCoV-19/England/CAMB-7ED51/2020|EPI_ISL_442898|202  .................... .................... .................... .................... .................... ....................
B  6883 hCoV-19/England/CAMB-7F752/2020|EPI_ISL_442899|202  .................... .................... .................... .................... .................... ....................
B  6884 hCoV-19/USA/FL-BPHL-0101/2020|EPI_ISL_445043|2020-  .................... .................... .................... .................... .................... ....................
B  6885 hCoV-19/Luxembourg/LNS0724201/2020|EPI_ISL_445054|  .................... .................... .................... .................... .................... ....................
B  6886 hCoV-19/Luxembourg/LNS5742995/2020|EPI_ISL_445055|  .................... .................... .................... .................... .................... ....................
B  6887 hCoV-19/Luxembourg/LNS9865959/2020|EPI_ISL_445056|  .................... .................... .................... .................... .................... ....................
B  6888 hCoV-19/Luxembourg/LNS8705424/2020|EPI_ISL_445057|  .................... .................... .................... .................... .................... ....................
B  6889 hCoV-19/Luxembourg/LNS9102463/2020|EPI_ISL_445058|  .................... .................... .................... .................... .................... ....................
B  6890 hCoV-19/Luxembourg/LNS6613781/2020|EPI_ISL_445060|  .................... .................... .................... .................... .................... ....................
B  6891 hCoV-19/Luxembourg/LNS1227733/2020|EPI_ISL_445061|  .................... .................... .................... .................... .................... ....................
B  6892 hCoV-19/Luxembourg/LNS0001029/2020|EPI_ISL_445064|  .................... .................... .................... .................... .................... ....................
B  6893 hCoV-19/Luxembourg/LNS1667297/2020|EPI_ISL_445065|  .................... .................... .................... .................... .................... ....................
B  6894 hCoV-19/Luxembourg/LNS2335124/2020|EPI_ISL_445066|  .................... .................... .................... .................... .................... ....................
B  6895 hCoV-19/Luxembourg/LNS9063347/2020|EPI_ISL_445067|  .................... .................... .................... .................... .................... ....................
B  6896 hCoV-19/Luxembourg/LNS3029697/2020|EPI_ISL_445069|  .................... .................... .................... .................... .................... ....................
B  6897 hCoV-19/USA/CA-SR0094/2020|EPI_ISL_445094|2020-03-  .................... .................... .................... .................... .................... ....................
B  6898 hCoV-19/USA/CA-SR0097/2020|EPI_ISL_445096|2020-03-  .................... .................... .................... .................... .................... ....................
B  6899 hCoV-19/USA/CA-SR0098/2020|EPI_ISL_445097|2020-03-  .................... .................... .................... .................... .................... ....................
B  6900 hCoV-19/USA/CA-SR0099/2020|EPI_ISL_445098|2020-03-  .................... .................... .................... .................... .................... ....................
B  6901 hCoV-19/Wales/PHWC-30E0D/2020|EPI_ISL_446363|2020-  .................... .................... .................... .................... .................... ....................
B  6902 hCoV-19/Wales/PHWC-30FA0/2020|EPI_ISL_446388|2020-  .................... ...A................ .................... .................... .................... ....................
B  6903 hCoV-19/France/IDF-10059IS/2020|EPI_ISL_447690|202  .................... .................... .................... .................... .................... ....................
B  6904 hCoV-19/France/IDF-10068ND/2020|EPI_ISL_447691|202  .................... .................... .................... .................... .................... ....................
B  6905 hCoV-19/France/IDF-40007RD/2020|EPI_ISL_447692|202  .................... .................... .................... .................... .................... ....................
B  6906 hCoV-19/France/IDF-5003KM/2020|EPI_ISL_447693|2020  .................... .................... .................... .................... .................... ....................
B  6907 hCoV-19/France/IDF-50002BS/2020|EPI_ISL_447694|202  .................... .................... .................... .................... .................... ....................
B  6908 hCoV-19/France/IDF-10004CF/2020|EPI_ISL_447695|202  .................... .................... .................... .................... .................... ....................
B  6909 hCoV-19/France/IDF-10009EE/2020|EPI_ISL_447696|202  .................... .................... .................... .................... .................... ....................
B  6910 hCoV-19/France/IDF-10008DM/2020|EPI_ISL_447697|202  .................... .................... .................... .................... .................... ....................
B  6911 hCoV-19/France/IDF-50001AR/2020|EPI_ISL_447698|202  .................... .................... .................... .................... .................... ....................
B  6912 hCoV-19/France/IDF-10010LA/2020|EPI_ISL_447699|202  .................... .................... .................... .................... .................... ....................
B  6913 hCoV-19/Argentina/PAIS-A0019/2020|EPI_ISL_430810|2  .................... .................... .................... .................... .................... ....................
B  6914 hCoV-19/Argentina/PAIS-A0020/2020|EPI_ISL_430811|2  .................... .................... .................... .................... .................... ....................
B  6915 hCoV-19/Argentina/PAIS-A0021/2020|EPI_ISL_430812|2  .................... .................... .................... .................... .................... ....................
B  6917 hCoV-19/Argentina/PAIS-A0023/2020|EPI_ISL_430814|2  .................... .................... .................... .................... .................... ....................
B  6918 hCoV-19/Argentina/PAIS-A0024/2020|EPI_ISL_430815|2  .................... .................... .................... .................... .................... ....................
B  6919 hCoV-19/Argentina/PAIS-A0025/2020|EPI_ISL_430816|2  .................... .................... .................... .................... .................... ....................
B  6920 hCoV-19/Argentina/PAIS-A0026/2020|EPI_ISL_430817|2  .................... .................... .................... .................... .................... ....................
B  6921 hCoV-19/Argentina/PAIS-A0027/2020|EPI_ISL_430818|2  .................... .................... .................... .................... .................... ....................
B  6922 hCoV-19/Egypt/NRC-03/2020|EPI_ISL_430819|2020-03-1  .................... .................... .................... .................... .................... ....................
B  6923 hCoV-19/Egypt/NRC-01/2020|EPI_ISL_430820|2020-03-1  .................... .................... .................... .................... .................... ....................
B  6924 hCoV-19/Thailand/NIH-2982/2020|EPI_ISL_430842|2020  .................... .................... .................... .................... .................... ....................
B  6925 hCoV-19/Sweden/20-04631/2020|EPI_ISL_430847|2020-0  .................... .................... .................... .................... .................... ....................
B  6926 hCoV-19/Sweden/20-08144/2020|EPI_ISL_430848|2020-0  .................... .................... .................... .................... .................... ....................
B  6927 hCoV-19/Sweden/20-08147/2020|EPI_ISL_430849|2020-0  .................... .................... .................... .................... .................... ....................
B  6928 hCoV-19/England/NOTT-10F3E9/2020|EPI_ISL_433000|20  .................... .................... .................... .................... .................... ....................
B  6929 hCoV-19/England/NOTT-10F404/2020|EPI_ISL_433001|20  .................... .................... .................... .................... .................... ....................
B  6930 hCoV-19/England/NOTT-10F413/2020|EPI_ISL_433002|20  .................... .................... .................... .................... .................... ....................
B  6931 hCoV-19/England/NOTT-10F4F5/2020|EPI_ISL_433012|20  .................... .................... .................... .................... .................... ....................
B  6932 hCoV-19/England/NOTT-10F53E/2020|EPI_ISL_433016|20  .................... .................... .................... .................... .................... ....................
```

| | | | | | | | | |
|---|---|---|---|---|---|---|---|---|
| B | 6933 hCoV-19/England/NOTT-10F695/2020\|EPI_ISL_433034\|20 | ···················· | ···················· | ···················· | ···················· | ···················· | ···················· | |
| B | 6934 hCoV-19/England/NOTT-10F792/2020\|EPI_ISL_433049\|20 | ···················· | ···················· | ···················· | ···················· | ···················· | ···················· | |
| B | 6940 hCoV-19/Scotland/EDB1631/2020\|EPI_ISL_433095\|2020- | ···················· | ···················· | ···················· | ···················· | ···················· | ···················· | |
| B | 6941 hCoV-19/Scotland/EDB1632/2020\|EPI_ISL_433096\|2020- | ···················· | ···················· | ···················· | ···················· | ···················· | ···················· | |
| B | 6943 hCoV-19/Scotland/EDB1638/2020\|EPI_ISL_433098\|2020- | ···················· | ···················· | ···················· | ···················· | ···················· | ···N·····NN········· | 1 |
| B | 6944 hCoV-19/Scotland/EDB1640/2020\|EPI_ISL_433099\|2020- | ···················· | ···················· | ···················· | ···················· | ···················· | ···················· | |
| B | 6945 hCoV-19/USA/WA-S540/2020\|EPI_ISL_434300\|2020-03-24 | ···················· | ···················· | ···················· | ···················· | ···················· | ···················· | |
| B | 6946 hCoV-19/USA/WA-S541/2020\|EPI_ISL_434301\|2020-03-24 | ···················· | ···················· | ···················· | ···················· | ···················· | ···················· | |
| B | 6947 hCoV-19/USA/WA-S542/2020\|EPI_ISL_434302\|2020-03-23 | ···················· | ···················· | ···················· | ···················· | ···················· | ···················· | |
| B | 6948 hCoV-19/USA/WA-S543/2020\|EPI_ISL_434303\|2020-03-24 | ···················· | ···················· | ···················· | ···················· | ···················· | ···················· | |
| B | 6949 hCoV-19/USA/WA-S545/2020\|EPI_ISL_434305\|2020-03-31 | ···················· | ···················· | ···················· | ···················· | ···················· | ···················· | |
| B | 6950 hCoV-19/USA/WA-S546/2020\|EPI_ISL_434306\|2020-03-23 | ···················· | ···················· | ···················· | ···················· | ···················· | ···················· | |
| B | 6951 hCoV-19/USA/WA-S547/2020\|EPI_ISL_434307\|2020-03-24 | ···················· | ···················· | ···················· | ···················· | ···················· | ···················· | |
| B | 6952 hCoV-19/USA/WA-S548/2020\|EPI_ISL_434308\|2020-03-31 | ···················· | ···················· | ···················· | ···················· | ···················· | ···················· | |
| B | 6953 hCoV-19/USA/WA-S549/2020\|EPI_ISL_434309\|2020-03-30 | ···················· | ···················· | ···················· | ···················· | ···················· | ···················· | |
| B | 6954 hCoV-19/USA/WA-S550/2020\|EPI_ISL_434310\|2020-03-24 | ···················· | ···················· | ···················· | ···················· | ···················· | ···················· | |
| B | 6955 hCoV-19/USA/WA-S551/2020\|EPI_ISL_434311\|2020-03-24 | ···················· | ···················· | ···················· | ···················· | ···················· | ···················· | |
| B | 6956 hCoV-19/USA/WA-S553/2020\|EPI_ISL_434313\|2020-03-31 | ···················· | ···················· | ···················· | ···················· | ···················· | ···················· | |
| B | 6957 hCoV-19/USA/WA-S554/2020\|EPI_ISL_434314\|2020-03-31 | ···················· | ···················· | ···················· | ···················· | ···················· | ···················· | |
| B | 6958 hCoV-19/USA/WA-S555/2020\|EPI_ISL_434315\|2020-03-30 | ···················· | ···················· | ···················· | ···················· | ···················· | ···················· | |
| B | 6959 hCoV-19/USA/WA-S556/2020\|EPI_ISL_434316\|2020-03-29 | ···················· | ···················· | ···················· | ···················· | ···················· | ···················· | |
| B | 6960 hCoV-19/USA/WA-S557/2020\|EPI_ISL_434317\|2020-04-01 | ···················· | ···················· | ···················· | ···················· | ···················· | ···················· | |
| B | 6961 hCoV-19/USA/WA-S559/2020\|EPI_ISL_434319\|2020-03-27 | ···················· | ···················· | ···················· | ···················· | ···················· | ···················· | |
| B | 6962 hCoV-19/USA/WA-S560/2020\|EPI_ISL_434320\|2020-03-24 | ···················· | ···················· | ···················· | ···················· | ···················· | ···················· | |
| B | 6963 hCoV-19/USA/WA-S561/2020\|EPI_ISL_434321\|2020-03-30 | ···················· | ···················· | ···················· | ···················· | ···················· | ···················· | |
| B | 6964 hCoV-19/USA/WA-S562/2020\|EPI_ISL_434322\|2020-03-24 | ···················· | ···················· | ···················· | ···················· | ···················· | ···················· | |
| B | 6965 hCoV-19/USA/WA-S563/2020\|EPI_ISL_434323\|2020-03-31 | ···················· | ···················· | ···················· | ···················· | ···················· | ···················· | |
| B | 6966 hCoV-19/USA/WA-S564/2020\|EPI_ISL_434324\|2020-03-24 | ···················· | ···················· | ···················· | ···················· | ···················· | ···················· | |
| B | 6967 hCoV-19/USA/WA-S567/2020\|EPI_ISL_434327\|2020-03-25 | ···················· | ···················· | ···················· | ···················· | ···················· | ···················· | |
| B | 6968 hCoV-19/USA/WA-S568/2020\|EPI_ISL_434328\|2020-03-31 | ···················· | ···················· | ···················· | ···················· | ···················· | ···················· | |
| B | 6969 hCoV-19/USA/WA-S569/2020\|EPI_ISL_434329\|2020-03-25 | ···················· | ···················· | ···················· | ···················· | ···················· | ···················· | |
| B | 6970 hCoV-19/USA/WA-S570/2020\|EPI_ISL_434330\|2020-03-24 | ···················· | ···················· | ···················· | ···················· | ···················· | ···················· | |
| B | 6971 hCoV-19/USA/WA-S571/2020\|EPI_ISL_434331\|2020-03-30 | ···················· | ···················· | ···················· | ···················· | ···················· | ···················· | |
| B | 6972 hCoV-19/USA/WA-S572/2020\|EPI_ISL_434332\|2020-03-26 | ···················· | ···················· | ···················· | ···················· | ···················· | ···················· | |
| B | 6973 hCoV-19/USA/WA-S574/2020\|EPI_ISL_434334\|2020-03-31 | ···················· | ···················· | ···················· | ···················· | ···················· | ···················· | |
| B | 6974 hCoV-19/USA/WA-S575/2020\|EPI_ISL_434335\|2020-03-31 | ···················· | ···················· | ···················· | ···················· | ···················· | ···················· | |
| B | 6975 hCoV-19/USA/WA-S576/2020\|EPI_ISL_434336\|2020-03-29 | ···················· | ···················· | ···················· | ···················· | ···················· | ···················· | |
| B | 6976 hCoV-19/USA/WA-S577/2020\|EPI_ISL_434337\|2020-03-28 | ···················· | ···················· | ···················· | ···················· | ···················· | ···················· | |
| B | 6977 hCoV-19/USA/WA-S578/2020\|EPI_ISL_434338\|2020-03-30 | ···················· | ···················· | ···················· | ···················· | ···················· | ···················· | |
| B | 6978 hCoV-19/USA/WA-S579/2020\|EPI_ISL_434339\|2020-03-24 | ···················· | ···················· | ···················· | ···················· | ···················· | ···················· | |
| B | 6979 hCoV-19/USA/WA-S581/2020\|EPI_ISL_434341\|2020-03-30 | ···················· | ···················· | ···················· | ···················· | ···················· | ···················· | |
| B | 6980 hCoV-19/USA/WA-S582/2020\|EPI_ISL_434342\|2020-03-31 | ···················· | ···················· | ···················· | ···················· | ···················· | ···················· | |
| B | 6981 hCoV-19/USA/WA-S583/2020\|EPI_ISL_434343\|2020-03-24 | ···················· | ···················· | ···················· | ···················· | ···················· | ···················· | |
| B | 6982 hCoV-19/USA/WA-S585/2020\|EPI_ISL_434345\|2020-03-26 | ···················· | ···················· | ···················· | ···················· | ···················· | ···················· | |
| B | 6983 hCoV-19/USA/WA-S586/2020\|EPI_ISL_434346\|2020-03-31 | ···················· | ···················· | ···················· | ···················· | ···················· | ···················· | |
| B | 6984 hCoV-19/Belgium/UGent-28/2020\|EPI_ISL_434347\|2020- | ···················· | ···················· | ···················· | ···················· | ···················· | ···················· | |
| B | 6985 hCoV-19/Belgium/UGent-44/2020\|EPI_ISL_434360\|2020- | ···················· | ···················· | ···················· | ···················· | ···················· | ···················· | |
| B | 6986 hCoV-19/Belgium/UGent-45/2020\|EPI_ISL_434361\|2020- | ···················· | ···················· | ···················· | ···················· | ···················· | ···················· | |
| B | 6987 hCoV-19/Belgium/UGent-46/2020\|EPI_ISL_434362\|2020- | ···················· | ···················· | ···················· | ···················· | ···················· | ···················· | |
| B | 6989 hCoV-19/Belgium/ITM_C211/2020\|EPI_ISL_434368\|2020- | ···················· | ···················· | ···················· | ···················· | ···················· | ···················· | |
| B | 6990 hCoV-19/Belgium/ITM_C262/2020\|EPI_ISL_434369\|2020- | ···················· | ···················· | ···················· | ···················· | ···················· | ···················· | |
| B | 6993 hCoV-19/Belgium/ITM_C053/2020\|EPI_ISL_434372\|2020- | ···················· | ···················· | ···················· | ···················· | ···················· | ···················· | |
| B | 6994 hCoV-19/Belgium/ITM_C147/2020\|EPI_ISL_434373\|2020- | ···················· | ···················· | ···················· | ···················· | ···················· | ···················· | |
| B | 7001 hCoV-19/USA/CA-SCCPHD-UC148/2020\|EPI_ISL_435627\|20 | ···················· | ···················· | ···················· | ···················· | ···················· | ···················· | |
| B | 7002 hCoV-19/USA/CA-SCCPHD-UC152/2020\|EPI_ISL_435631\|20 | ···················· | ···················· | ···················· | ···················· | ···················· | ···················· | |
| B | 7003 hCoV-19/USA/CA-SCCPHD-UC155/2020\|EPI_ISL_435634\|20 | ···················· | ···················· | ···················· | ···················· | ···················· | ···················· | |
| B | 7004 hCoV-19/USA/CA-SCCPHD-UC157/2020\|EPI_ISL_435636\|20 | ···················· | ···················· | ···················· | ···················· | ···················· | ···················· | |
| B | 7005 hCoV-19/USA/CA-SCCPHD-UC158/2020\|EPI_ISL_435637\|20 | ···················· | ···················· | ···················· | ···················· | ···················· | ···················· | |
| B | 7006 hCoV-19/USA/CA-SCCPHD-UC160/2020\|EPI_ISL_435639\|20 | ···················· | ···················· | ···················· | ···················· | ···················· | ···················· | |
| B | 7007 hCoV-19/USA/CA-SCCPHD-UC162/2020\|EPI_ISL_435641\|20 | ···················· | ···················· | ···················· | ···················· | ···················· | ···················· | |
| B | 7008 hCoV-19/USA/CA-SCCPHD-UC164/2020\|EPI_ISL_435643\|20 | ···················· | ···················· | ···················· | ···················· | ···················· | ···················· | |
| B | 7009 hCoV-19/USA/CA-SCCPHD-UC166/2020\|EPI_ISL_435645\|20 | ···················· | ···················· | ···················· | ···················· | ···················· | ···················· | |
| B | 7010 hCoV-19/USA/CA-SCCPHD-UC170/2020\|EPI_ISL_435649\|20 | ···················· | ···················· | ···················· | ···················· | ···················· | ···················· | |
| B | 7011 hCoV-19/USA/CA-SCCPHD-UC171/2020\|EPI_ISL_435650\|20 | ···················· | ···················· | ···················· | ···················· | ···················· | ···················· | |
| B | 7012 hCoV-19/USA/CA-SCCPHD-UC175/2020\|EPI_ISL_435654\|20 | ···················· | ···················· | ···················· | ···················· | ···················· | ···················· | |
| B | 7014 hCoV-19/USA/CA-SCCPHD-UC183/2020\|EPI_ISL_435662\|20 | ···················· | ···················· | ···················· | ···················· | ···················· | NNN················· | 1 |
| B | 7015 hCoV-19/USA/CA-SCCPHD-UC184/2020\|EPI_ISL_435663\|20 | ···················· | ···················· | ···················· | ···················· | ···················· | ···················· | |
| B | 7016 hCoV-19/USA/CA-SCCPHD-UC191/2020\|EPI_ISL_435670\|20 | ···················· | ···················· | ···················· | ···················· | ···················· | ···················· | |
| B | 7017 hCoV-19/USA/CA-SCCPHD-UC192/2020\|EPI_ISL_435671\|20 | ···················· | ···················· | ···················· | ···················· | ···················· | ···················· | |
| B | 7018 hCoV-19/Brunei/2/2020\|EPI_ISL_435674\|2020-03-11 | ···················· | ···················· | ···················· | ···················· | ···················· | ···················· | |
| B | 7019 hCoV-19/Brunei/3/2020\|EPI_ISL_435675\|2020-03-11 | ···················· | ···················· | ···················· | ···················· | ···················· | ···················· | |
| B | 7020 hCoV-19/Brunei/4/2020\|EPI_ISL_435676\|2020-03-21 | ···················· | ···················· | ···················· | ···················· | ···················· | ···················· | |
| B | 7021 hCoV-19/Brunei/5/2020\|EPI_ISL_435677\|2020-03-21 | ···················· | ···················· | ···················· | ···················· | ···················· | ···················· | |
| B | 7022 hCoV-19/Singapore/73/2020\|EPI_ISL_435678\|2020-03-2 | ···················· | ···················· | ···················· | ···················· | ···················· | ···················· | |
| B | 7023 hCoV-19/Singapore/85/2020\|EPI_ISL_435690\|2020-04-1 | ···················· | ···················· | ···················· | ···················· | ···················· | ···················· | |
| B | 7024 hCoV-19/Singapore/86/2020\|EPI_ISL_435691\|2020-04-2 | ···················· | ···················· | ···················· | ···················· | ···················· | ···················· | |
| B | 7025 hCoV-19/Singapore/87/2020\|EPI_ISL_435692\|2020-04-2 | ···················· | ···················· | ···················· | ···················· | ···················· | ···················· | |
| B | 7026 hCoV-19/Singapore/88/2020\|EPI_ISL_435693\|2020-04-2 | ···················· | ···················· | ···················· | ···················· | ···················· | ···················· | |

| | | | | | | | | | |
|---|---|---|---|---|---|---|---|---|---|
| B | 7027 | hCoV-19/Singapore/89/2020\|EPI_ISL_435694\|2020-04-2 | .................... | .................... | .................... | .................... | .................... | .................... | |
| B | 7028 | hCoV-19/Singapore/90/2020\|EPI_ISL_435695\|2020-04-2 | .................... | .................... | .................... | .................... | .................... | .................... | |
| B | 7029 | hCoV-19/Singapore/91/2020\|EPI_ISL_435696\|2020-04-2 | .................... | .................... | .................... | .................... | .................... | .................... | |
| B | 7030 | hCoV-19/Singapore/92/2020\|EPI_ISL_435697\|2020-03-1 | .................... | .................... | .................... | .................... | .................... | .................... | |
| B | 7031 | hCoV-19/Singapore/93/2020\|EPI_ISL_435698\|2020-03-1 | .................... | .................... | .................... | .................... | .................... | .................... | |
| B | 7032 | hCoV-19/Singapore/94/2020\|EPI_ISL_435699\|2020-03-1 | .................... | .................... | .................... | .................... | .................... | .................... | |
| B | 7033 | hCoV-19/USA/LA-BIE-088/2020\|EPI_ISL_436956\|2020-04 | .................... | .................... | .................... | .................... | .................... | .................... | |
| B | 7034 | hCoV-19/USA/LA-BIE-089/2020\|EPI_ISL_436957\|2020-04 | .................... | .................... | .................... | .................... | .................... | .................... | |
| B | 7035 | hCoV-19/USA/LA-BIE-090/2020\|EPI_ISL_436958\|2020-04 | .................... | .................... | .................... | .................... | .................... | .................... | |
| B | 7036 | hCoV-19/USA/LA-BIE-092/2020\|EPI_ISL_436959\|2020-04 | .................... | .................... | .................... | .................... | .................... | .................... | |
| B | 7037 | hCoV-19/USA/LA-BIE-093/2020\|EPI_ISL_436960\|2020-04 | .................... | .................... | .................... | .................... | .................... | .................... | |
| B | 7038 | hCoV-19/USA/LA-BIE-094/2020\|EPI_ISL_436961\|2020-04 | .................... | .................... | .................... | .................... | .................... | .................... | |
| B | 7039 | hCoV-19/Denmark/ALAB-SSI-1126/2020\|EPI_ISL_436962\| | .................... | .................... | .................... | .................... | .................... | .................... | |
| B | 7040 | hCoV-19/Denmark/ALAB-SSI-197/2020\|EPI_ISL_436963\|2 | .................... | .................... | .................... | .................... | .................... | .................... | |
| B | 7041 | hCoV-19/Denmark/ALAB-SSI-284/2020\|EPI_ISL_436965\|2 | .................... | .................... | .................... | .................... | .................... | .................... | |
| B | 7042 | hCoV-19/Denmark/ALAB-SSI-547/2020\|EPI_ISL_436967\|2 | .................... | .................... | .................... | .................... | .................... | .................... | |
| B | 7043 | hCoV-19/Denmark/ALAB-SSI-559/2020\|EPI_ISL_436969\|2 | .................... | .................... | .................... | .................... | .................... | .................... | |
| B | 7044 | hCoV-19/Denmark/ALAB-SSI-570/2020\|EPI_ISL_436973\|2 | .................... | .................... | .................... | .................... | .................... | .................... | |
| B | 7045 | hCoV-19/Denmark/ALAB-SSI-572/2020\|EPI_ISL_436975\|2 | .................... | .................... | .................... | .................... | .................... | .................... | |
| B | 7046 | hCoV-19/Denmark/ALAB-SSI-579/2020\|EPI_ISL_436978\|2 | .................... | ...................A | .................... | .................... | .................... | .................... | 1 |
| B | 7047 | hCoV-19/Denmark/ALAB-SSI-580/2020\|EPI_ISL_436979\|2 | .................... | ...................N | .................... | .................... | .................... | .................... | 1 |
| B | 7048 | hCoV-19/Denmark/ALAB-SSI-581/2020\|EPI_ISL_436980\|2 | .................... | .................... | .................... | .................... | .................... | .................... | |
| B | 7049 | hCoV-19/Denmark/ALAB-SSI-582/2020\|EPI_ISL_436981\|2 | .................... | ...................A | .................... | .................... | .................... | .................... | 1 |
| B | 7050 | hCoV-19/Denmark/ALAB-SSI-586/2020\|EPI_ISL_436982\|2 | .................... | .................... | .................... | .................... | .................... | .................... | |
| B | 7051 | hCoV-19/Denmark/ALAB-SSI-590/2020\|EPI_ISL_436984\|2 | .................... | .................... | .................... | .................... | .................... | .................... | |
| B | 7052 | hCoV-19/Denmark/ALAB-SSI-596/2020\|EPI_ISL_436985\|2 | .................... | .................... | .................... | .................... | .................... | .................... | |
| B | 7053 | hCoV-19/Denmark/ALAB-SSI-598/2020\|EPI_ISL_436986\|2 | .................... | .................... | .................... | .................... | .................... | .................... | |
| B | 7054 | hCoV-19/Denmark/ALAB-SSI-600/2020\|EPI_ISL_436987\|2 | .................... | .................... | .................... | .................... | .................... | .................... | |
| B | 7055 | hCoV-19/Denmark/ALAB-SSI-604/2020\|EPI_ISL_436988\|2 | .................... | .................... | .................... | .................... | .................... | .................... | |
| B | 7056 | hCoV-19/Denmark/ALAB-SSI-605/2020\|EPI_ISL_436989\|2 | .................... | .................... | .................... | .................... | .................... | .................... | |
| B | 7057 | hCoV-19/Denmark/ALAB-SSI-607/2020\|EPI_ISL_436990\|2 | .................... | .................... | .................... | .................... | .................... | .................... | |
| B | 7058 | hCoV-19/Denmark/ALAB-SSI-608/2020\|EPI_ISL_436991\|2 | .................... | .................... | .................... | .................... | .................... | .................... | |
| B | 7059 | hCoV-19/Denmark/ALAB-SSI-611/2020\|EPI_ISL_436993\|2 | .................... | .................... | .................... | .................... | .................... | .................... | |
| B | 7060 | hCoV-19/Denmark/ALAB-SSI-613/2020\|EPI_ISL_436995\|2 | .................... | .................... | .................... | .................... | .................... | .................... | |
| B | 7061 | hCoV-19/Denmark/ALAB-SSI-614/2020\|EPI_ISL_436996\|2 | .................... | .................... | .................... | .................... | .................... | .................... | |
| B | 7062 | hCoV-19/Denmark/ALAB-SSI-615/2020\|EPI_ISL_436997\|2 | .................... | .................... | .................... | .................... | .................... | .................... | |
| B | 7063 | hCoV-19/Denmark/ALAB-SSI-616/2020\|EPI_ISL_436998\|2 | .................... | .................... | .................... | .................... | .................... | .................... | |
| B | 7064 | hCoV-19/Denmark/ALAB-SSI-617/2020\|EPI_ISL_436999\|2 | .................... | .................... | .................... | .................... | .................... | .................... | |
| B | 7065 | hCoV-19/England/BRIS-123ABD/2020\|EPI_ISL_442800\|20 | .................... | .................... | .................... | .................... | .................... | .................... | |
| B | 7066 | hCoV-19/England/BRIS-123534/2020\|EPI_ISL_442801\|20 | .................... | .................... | .................... | .................... | .................... | .................... | |
| B | 7067 | hCoV-19/England/BRIS-123B6F/2020\|EPI_ISL_442802\|20 | .................... | .................... | .................... | .................... | .................... | .................... | |
| B | 7068 | hCoV-19/England/BRIS-12332B/2020\|EPI_ISL_442804\|20 | .................... | .................... | .................... | .................... | .................... | .................... | |
| B | 7069 | hCoV-19/England/BRIS-123BAB/2020\|EPI_ISL_442805\|20 | .................... | .................... | .................... | .................... | .................... | .................... | |
| B | 7070 | hCoV-19/England/BRIS-123A71/2020\|EPI_ISL_442807\|20 | .................... | .................... | .................... | .................... | .................... | .................... | |
| B | 7071 | hCoV-19/England/BRIS-123895/2020\|EPI_ISL_442808\|20 | .................... | .................... | .................... | .................... | .................... | .................... | |
| B | 7072 | hCoV-19/England/BRIS-123060/2020\|EPI_ISL_442809\|20 | .................... | .................... | .................... | .................... | .................... | .................... | |
| B | 7073 | hCoV-19/England/CAMB-7BFB8/2020\|EPI_ISL_442820\|202 | .................... | .................... | .................... | .................... | .................... | .................... | |
| B | 7074 | hCoV-19/England/CAMB-81614/2020\|EPI_ISL_442822\|202 | .................... | .................... | .................... | .................... | .................... | .................... | |
| B | 7075 | hCoV-19/England/CAMB-81289/2020\|EPI_ISL_442823\|202 | .................... | .................... | .................... | .................... | .................... | .................... | |
| B | 7076 | hCoV-19/England/CAMB-7EDBB/2020\|EPI_ISL_442824\|202 | .................... | .................... | .................... | .................... | .................... | .................... | |
| B | 7077 | hCoV-19/England/CAMB-7F0A2/2020\|EPI_ISL_442825\|202 | .................... | .................... | .................... | .................... | .................... | .................... | |
| B | 7078 | hCoV-19/England/CAMB-7EC81/2020\|EPI_ISL_442826\|202 | .................... | .................... | .................... | .................... | .................... | .................... | |
| B | 7079 | hCoV-19/England/CAMB-80DB6/2020\|EPI_ISL_442828\|202 | .................... | .................... | .................... | .................... | .................... | .................... | |
| B | 7080 | hCoV-19/England/CAMB-7EED6/2020\|EPI_ISL_442829\|202 | .................... | .................... | .................... | .................... | .................... | .................... | |
| B | 7081 | hCoV-19/Luxembourg/LNS0857930/2020\|EPI_ISL_445070\| | .................... | .................... | .................... | .................... | .................... | .................... | |
| B | 7082 | hCoV-19/Luxembourg/LNS7188652/2020\|EPI_ISL_445071\| | .................... | .................... | .................... | .................... | .................... | .................... | |
| B | 7083 | hCoV-19/Luxembourg/LNS5711030/2020\|EPI_ISL_445072\| | .................... | .................... | .................... | .................... | .................... | .................... | |
| B | 7084 | hCoV-19/Luxembourg/LNS2156711/2020\|EPI_ISL_445074\| | .................... | .................... | .................... | .................... | .................... | .................... | |
| B | 7085 | hCoV-19/Luxembourg/LNS1586475/2020\|EPI_ISL_445075\| | .................... | .................... | .................... | .................... | .................... | .................... | |
| B | 7086 | hCoV-19/Luxembourg/LNS7550599/2020\|EPI_ISL_445076\| | .................... | .................... | .................... | .................... | .................... | .................... | |
| B | 7087 | hCoV-19/USA/MN-UMGC-7/2020\|EPI_ISL_445077\|2020-03- | .................... | .................... | .................... | .................... | .................... | .................... | |
| B | 7088 | hCoV-19/USA/TX_GCID_192000021/2020\|EPI_ISL_445078\| | .................... | .................... | .................... | .................... | .................... | .................... | |
| B | 7089 | hCoV-19/USA/TX_GCID_192000052/2020\|EPI_ISL_445079\| | .................... | .................... | .................... | .................... | .................... | .................... | |
| B | 7090 | hCoV-19/USA/TX_GCID_192000072/2020\|EPI_ISL_445080\| | .................... | .................... | .................... | .................... | .................... | .................... | |
| B | 7091 | hCoV-19/USA/TX_GCID_192000088/2020\|EPI_ISL_445081\| | .................... | .................... | .................... | .................... | .................... | .................... | |
| B | 7092 | hCoV-19/USA/TX_GCID_192000119/2020\|EPI_ISL_445082\| | .................... | .................... | .................... | .................... | .................... | .................... | |
| B | 7093 | hCoV-19/USA/TX_GCID_192000124/2020\|EPI_ISL_445083\| | .................... | .................... | .................... | .................... | .................... | .................... | |
| B | 7094 | hCoV-19/USA/TX_GCID_192000135/2020\|EPI_ISL_445084\| | .................... | .................... | .................... | .................... | .................... | .................... | |
| B | 7095 | hCoV-19/Colombia/un-CIAT-01/2020\|EPI_ISL_445085\|20 | .................... | .................... | .................... | .................... | .................... | .................... | |
| B | 7096 | hCoV-19/Serbia/KV26/2020\|EPI_ISL_445086\|2020-04-13 | .................... | .................... | .................... | .................... | .................... | .................... | |
| B | 7097 | hCoV-19/Serbia/Novi Pazar-363/2020\|EPI_ISL_445087\| | .................... | .................... | .................... | .................... | .................... | .................... | |
| B | 7098 | hCoV-19/Wales/PHWC-309A2/2020\|EPI_ISL_446301\|2020- | .................... | .................... | .................... | .................... | .................... | .................... | |
| B | 7099 | hCoV-19/Wales/PHWC-309FD/2020\|EPI_ISL_446305\|2020- | .................... | .................... | .................... | .................... | .................... | .................... | |
| B | 7100 | hCoV-19/Wales/PHWC-30A18/2020\|EPI_ISL_446307\|2020- | .................... | ....A............... | .................... | .................... | .................... | .................... | |
| B | 7101 | hCoV-19/Wales/PHWC-30A36/2020\|EPI_ISL_446309\|2020- | .................... | .................... | .................... | .................... | .................... | .................... | |
| B | 7102 | hCoV-19/Wales/PHWC-30A45/2020\|EPI_ISL_446310\|2020- | .................... | .................... | .................... | .................... | .................... | .................... | |
| B | 7103 | hCoV-19/Wales/PHWC-30AEB/2020\|EPI_ISL_446320\|2020- | .................... | .................... | .................... | .................... | .................... | .................... | |
| B | 7104 | hCoV-19/Wales/PHWC-30B06/2020\|EPI_ISL_446321\|2020- | .................... | .................... | .................... | .................... | .................... | .................... | |

B 7105 hCoV-19/Wales/PHWC-30C21/2020|EPI_ISL_446339|2020- ···················· ···················· ···················· ···················· ···················· ····················
B 7106 hCoV-19/Wales/PHWC-30C30/2020|EPI_ISL_446340|2020- ···················· ···················· ···················· ···················· ···················· ····················
B 7107 hCoV-19/Wales/PHWC-30C7C/2020|EPI_ISL_446343|2020- ···················· ···················· ···················· ···················· ···················· ····················
B 7108 hCoV-19/Wales/PHWC-30CB8/2020|EPI_ISL_446347|2020- ···················· ···················· ···················· ···················· ···················· ····················
B 7109 hCoV-19/Wales/PHWC-30D00/2020|EPI_ISL_446352|2020- ···················· ···················· ···················· ···················· ···················· ····················
B 7110 hCoV-19/Germany/HE-FFM1clin/2020|EPI_ISL_447608|20 ···················· ·A·················· ···················· ···················· ···················· ····················
B 7111 hCoV-19/Germany/HE-FFM3clin/2020|EPI_ISL_447609|20 ···················· ···················· ···················· ···················· ···················· ····················
B 7112 hCoV-19/Germany/HE-FFM4clin/2020|EPI_ISL_447610|20 ···················· ···················· ···················· ···················· ···················· ····················
B 7113 hCoV-19/Germany/HE-FFM5clin/2020|EPI_ISL_447611|20 ···················· ···················· ···················· ···················· ···················· ····················
B 7114 hCoV-19/Germany/HE-FFM6clin/2020|EPI_ISL_447612|20 ···················· ···················· ···················· ···················· ···················· ····················
B 7115 hCoV-19/Germany/HE-FFM7clin/2020|EPI_ISL_447613|20 ···················· ···················· ···················· ···················· ···················· ····················
B 7116 hCoV-19/mink/Netherlands/NB01_02KS/2020|EPI_ISL_44 ···················· ···················· ···················· ···················· ···················· ····················
B 7117 hCoV-19/mink/Netherlands/NB01_03KS/2020|EPI_ISL_44 ···················· ···················· ···················· ···················· ···················· ····················
B 7118 hCoV-19/mink/Netherlands/NB01_04KS/2020|EPI_ISL_44 ···················· ···················· ···················· ···················· ···················· ····················
B 7119 hCoV-19/mink/Netherlands/NB02_03KS/2020|EPI_ISL_44 ···················· ···················· ···················· ···················· ····A··············· ····················
B 7120 hCoV-19/mink/Netherlands/NB02_06KS/2020|EPI_ISL_44 ···················· ···················· ···················· ···················· ····A··············· ····················
B 7121 hCoV-19/mink/Netherlands/NB02_07KS/2020|EPI_ISL_44 ···················· ···················· ···················· ···················· ···················· ····················
B 7122 hCoV-19/mink/Netherlands/NB02_13KS/2020|EPI_ISL_44 ···················· ···················· ···················· ···················· ···················· ····················
B 7123 hCoV-19/mink/Netherlands/NB02_index/2020|EPI_ISL_4 ···················· ···················· ···················· ···················· ····A··············· ····················
B 7124 hCoV-19/mink/Netherlands/NB02_16RS/2020|EPI_ISL_44 ···················· ···················· ···················· ···················· ····A··············· ····················
B 7125 hCoV-19/mink/Netherlands/NB03_index/2020|EPI_ISL_4 ···················· ···················· ···················· ···················· ···················· ····················
B 7126 hCoV-19/mink/Netherlands/NB04_index/2020|EPI_ISL_4 ···················· ···················· ···················· ···················· ···················· ····················
B 7127 hCoV-19/Greece/149_32357/2020|EPI_ISL_447635|2020- ···················· ···················· ···················· ···················· ···················· ····················
B 7128 hCoV-19/Greece/203_33026/2020|EPI_ISL_447636|2020- ···················· ···················· ···················· ···················· ···················· ····················
B 7129 hCoV-19/Greece/205_33490/2020|EPI_ISL_447637|2020- ···················· ···················· ···················· ···················· ···················· ····················
B 7130 hCoV-19/Greece/206_33492/2020|EPI_ISL_447638|2020- ···················· ···················· ···················· ···················· ···················· ····················
B 7131 hCoV-19/Greece/207_33548/2020|EPI_ISL_447639|2020- ···················· ···················· ···················· ···················· ···················· ····················
B 7132 hCoV-19/Greece/211_34005/2020|EPI_ISL_447640|2020- ···················· ···················· ···················· ···················· ···················· ····················
B 7133 hCoV-19/Greece/216_34732/2020|EPI_ISL_447641|2020- ···················· ···················· ···················· ···················· ···················· ····················
B 7134 hCoV-19/Greece/219_35198/2020|EPI_ISL_447642|2020- ···················· ···················· ···················· ···················· ···················· ····················
B 7135 hCoV-19/Greece/222_33921/2020|EPI_ISL_447643|2020- ···················· ···················· ···················· ···················· ···················· ····················
B 7136 hCoV-19/Greece/227_35969/2020|EPI_ISL_447644|2020- ···················· ···················· ···················· ···················· ···················· ····················
B 7137 hCoV-19/Greece/238_31927/2020|EPI_ISL_447645|2020- ···················· ···················· ···················· ···················· ···················· ····················
B 7138 hCoV-19/Greece/239_32178/2020|EPI_ISL_447646|2020- ···················· ···················· ···················· ···················· ···················· ····················
B 7139 hCoV-19/Greece/250_32267/2020|EPI_ISL_447648|2020- ···················· ···················· ···················· ···················· ···················· ····················
B 7140 hCoV-19/Greece/255_32525/2020|EPI_ISL_447650|2020- ···················· ···················· ···················· ···················· ···················· ····················
B 7141 hCoV-19/Greece/259_31928/2020|EPI_ISL_447651|2020- ···················· ···················· ···················· ···················· ···················· ····················
B 7142 hCoV-19/Greece/262_32443/2020|EPI_ISL_447652|2020- ···················· ···················· ···················· ···················· ···················· ····················
B 7143 hCoV-19/Greece/263_32477/2020|EPI_ISL_447653|2020- ···················· ···················· ···················· ···················· ···················· ····················
B 7144 hCoV-19/France/IDF-10027AA/2020|EPI_ISL_447654|202 ···················· ···················· ···················· ···················· ···················· ····················
B 7145 hCoV-19/France/IDF-70001GR/2020|EPI_ISL_447655|202 ···················· ···················· ···················· ···················· ···················· ····················
B 7146 hCoV-19/France/40004SR/2020|EPI_ISL_447656|2020-03 ···················· ···················· ···················· ···················· ···················· ····················
B 7147 hCoV-19/France/IDF-50010DC/2020|EPI_ISL_447657|202 ···················· ···················· ···················· ···················· ···················· ····················
B 7148 hCoV-19/France/IDF-10029DT/2020|EPI_ISL_447669|202 ···················· ···················· ···················· ···················· ···················· ····················
B 7149 hCoV-19/France/IDF-10031BS/2020|EPI_ISL_447670|202 ···················· ···················· ···················· ···················· ···················· ····················
B 7150 hCoV-19/France/IDF-60002VV/2020|EPI_ISL_447671|202 ···················· ···················· ···················· ···················· ···················· ····················
B 7151 hCoV-19/France/IDF-10032KL/2020|EPI_ISL_447672|202 ···················· ···················· ···················· ···················· ···················· ····················
B 7152 hCoV-19/France/IDF-50008KC/2020|EPI_ISL_447673|202 ···················· ···················· ···················· ···················· ···················· ····················
B 7153 hCoV-19/France/IDF-10033ZM/2020|EPI_ISL_447674|202 ···················· ···················· ···················· ···················· ···················· ····················
B 7154 hCoV-19/France/IDF-10034HM/2020|EPI_ISL_447675|202 ···················· ···················· ···················· ···················· ···················· ····················
B 7155 hCoV-19/France/IDF-10050RA/2020|EPI_ISL_447676|202 ···················· ···················· ···················· ···················· ···················· ····················
B 7156 hCoV-19/France/IDF-10045DZ/2020|EPI_ISL_447677|202 ···················· ···················· ···················· ···················· ···················· ····················
B 7157 hCoV-19/France/IDF-10051SM/2020|EPI_ISL_447678|202 ···················· ···················· ···················· ···················· ···················· ····················
B 7158 hCoV-19/France/IDF-10063BI/2020|EPI_ISL_447679|202 ···················· ···················· ···················· ···················· ···················· ····················
B 7159 hCoV-19/France/IDF-10066GJ/2020|EPI_ISL_447680|202 ···················· ···················· ···················· ···················· ···················· ····················
B 7160 hCoV-19/France/IDF-10069NM/2020|EPI_ISL_447681|202 ···················· ···················· ···················· ···················· ···················· ····················
B 7161 hCoV-19/France/IDF-10071BR/2020|EPI_ISL_447682|202 ···················· ···················· ···················· ···················· ···················· ····················
B 7162 hCoV-19/France/IDF-10060KV/2020|EPI_ISL_447683|202 ···················· ···················· ···················· ···················· ···················· ····················
B 7163 hCoV-19/France/IDF-10074LD/2020|EPI_ISL_447684|202 ···················· ···················· ···················· ···················· ···················· ····················
B 7164 hCoV-19/France/IDF-10072CG/2020|EPI_ISL_447685|202 ···················· ···················· ···················· ···················· ···················· ····················
B 7165 hCoV-19/France/IDF-10076MH/2020|EPI_ISL_447686|202 ···················· ···················· ···················· ···················· ···················· ····················
B 7166 hCoV-19/France/IDF-10078MA/2020|EPI_ISL_447687|202 ···················· ···················· ···················· ···················· ···················· ····················
B 7167 hCoV-19/France/IDF-10057CJ/2020|EPI_ISL_447688|202 ···················· ···················· ···················· ···················· ···················· ····················
B 7168 hCoV-19/France/IDF-10062MD/2020|EPI_ISL_447689|202 ···················· ···················· ···················· ···················· ···················· ····················
B 7169 hCoV-19/DRC/1319/2020|EPI_ISL_431011|2020-04-07 ···················· ···················· ···················· ···················· ···················· ····················
B 7170 hCoV-19/DRC/1376/2020|EPI_ISL_431012|2020-04-07 ···················· ···················· ···················· ···················· ···················· ····················
B 7171 hCoV-19/USA/AK-PHL010/2020|EPI_ISL_431013|2020-03- ···················· ···················· ···················· ···················· ···················· ····················
B 7172 hCoV-19/USA/AK-PHL011/2020|EPI_ISL_431014|2020-03- ···················· ···················· ···················· ···················· ···················· ····················
B 7173 hCoV-19/USA/AK-PHL013/2020|EPI_ISL_431015|2020-03- ···················· ···················· ···················· ···················· ···················· ····················
B 7174 hCoV-19/USA/AK-PHL016/2020|EPI_ISL_431016|2020-03- ···················· ···················· ···················· ···················· ···················· ····················
B 7175 hCoV-19/USA/AK-PHL040/2020|EPI_ISL_431017|2020-03- ···················· ···················· ···················· ···················· ···················· ····················
B 7176 hCoV-19/USA/AK-PHL052/2020|EPI_ISL_431018|2020-03- ···················· ···················· ···················· ···················· ···················· ····················
B 7177 hCoV-19/USA/AK-PHL074/2020|EPI_ISL_431019|2020-03- ···················· ···················· ···················· ···················· ···················· ····················
B 7178 hCoV-19/Wales/PHWC-2B096/2020|EPI_ISL_432300|2020- ···················· ···················· ···················· ···················· ···················· ····················
B 7179 hCoV-19/Wales/PHWC-27407/2020|EPI_ISL_432301|2020- ···················· ···················· ···················· ···················· ···················· ····················
B 7180 hCoV-19/Wales/PHWC-273A0/2020|EPI_ISL_432302|2020- ···················· ···················· ···················· ···················· ···················· ····················
B 7181 hCoV-19/Wales/PHWC-27759/2020|EPI_ISL_432303|2020- ···················· ···················· ···················· ···················· ···················· ····················
B 7182 hCoV-19/Wales/PHWC-2B157/2020|EPI_ISL_432304|2020- ···················· ···················· ···················· ···················· ···················· ····················

| | | | | | | | | |
|---|---|---|---|---|---|---|---|---|
| B | 7183 hCoV-19/Wales/PHWC-2B087/2020\|EPI_ISL_432305\|2020- | ···················· | ···················· | ····················· | ···················· | ···················· | ···················· | |
| B | 7184 hCoV-19/Wales/PHWC-27689/2020\|EPI_ISL_432306\|2020- | ···················· | ···················· | ····················· | ···················· | ···················· | ···················· | |
| B | 7185 hCoV-19/Wales/PHWC-27267/2020\|EPI_ISL_432307\|2020- | ···················· | ···················· | ····················· | ···················· | ···················· | ···················· | |
| B | 7186 hCoV-19/Wales/PHWC-24E39/2020\|EPI_ISL_432308\|2020- | ···················· | ···················· | ····················· | ···················· | ···················· | ···················· | |
| B | 7187 hCoV-19/Wales/PHWC-2B0A5/2020\|EPI_ISL_432309\|2020- | ···················· | ···················· | ····················· | ···················· | ···················· | ···················· | |
| B | 7188 hCoV-19/Wales/PHWC-27BB7/2020\|EPI_ISL_432310\|2020- | ···················· | ···················· | ····················· | ···················· | ···················· | ···················· | |
| B | 7189 hCoV-19/Wales/PHWC-26B6D/2020\|EPI_ISL_432311\|2020- | ···················· | ···················· | ····················· | ···················· | ···················· | ···················· | |
| B | 7190 hCoV-19/Wales/PHWC-271F1/2020\|EPI_ISL_432312\|2020- | ···················· | ···················· | ····················· | ···················· | ···················· | ···················· | |
| B | 7191 hCoV-19/Wales/PHWC-29FC8/2020\|EPI_ISL_432313\|2020- | ···················· | ···················· | ····················· | ···················· | ···················· | ···················· | |
| B | 7192 hCoV-19/Wales/PHWC-276B6/2020\|EPI_ISL_432314\|2020- | ···················· | ···················· | ····················· | ···················· | ···················· | ···················· | |
| B | 7193 hCoV-19/Wales/PHWC-2B2EB/2020\|EPI_ISL_432315\|2020- | ···················· | ···················· | ···A················· | ···················· | ···················· | ···················· | |
| B | 7194 hCoV-19/Wales/PHWC-274CB/2020\|EPI_ISL_432316\|2020- | ···················· | ···················· | ····················· | ···················· | ···················· | ···················· | |
| B | 7195 hCoV-19/Wales/PHWC-2723A/2020\|EPI_ISL_432317\|2020- | ···················· | ···················· | ····················· | ···················· | ···················· | ···················· | |
| B | 7196 hCoV-19/Wales/PHWC-2B290/2020\|EPI_ISL_432318\|2020- | ···················· | ····A··············· | ····················· | ···················· | ···················· | ···················· | |
| B | 7197 hCoV-19/Wales/PHWC-29EDA/2020\|EPI_ISL_432319\|2020- | ···················· | ···················· | ····················· | ···················· | ···················· | ···················· | |
| B | 7198 hCoV-19/Wales/PHWC-2ADCD/2020\|EPI_ISL_432320\|2020- | ···················· | ···················· | ····················· | ···················· | ···················· | ···················· | |
| B | 7199 hCoV-19/Wales/PHWC-27B02/2020\|EPI_ISL_432321\|2020- | ···················· | ···················· | ····················· | ···················· | ···················· | ···················· | |
| B | 7200 hCoV-19/Wales/PHWC-2780B/2020\|EPI_ISL_432322\|2020- | ···················· | ···················· | ····················· | ···················· | ···················· | ···················· | |
| B | 7201 hCoV-19/Wales/PHWC-25348/2020\|EPI_ISL_432323\|2020- | ···················· | ····A··············· | ····················· | ···················· | ···················· | ···················· | |
| B | 7202 hCoV-19/Wales/PHWC-27B6C/2020\|EPI_ISL_432325\|2020- | ···················· | ····A··············· | ····················· | ···················· | ···················· | ···················· | |
| B | 7203 hCoV-19/Wales/PHWC-247E3/2020\|EPI_ISL_432326\|2020- | ···················· | ···················· | ····················· | ···················· | ···················· | ···················· | |
| B | 7204 hCoV-19/Wales/PHWC-27B5D/2020\|EPI_ISL_432327\|2020- | ···················· | ···················· | ····················· | ···················· | ···················· | ···················· | |
| B | 7205 hCoV-19/Wales/PHWC-2756E/2020\|EPI_ISL_432328\|2020- | ···················· | ···················· | ····················· | ···················· | ···················· | ···················· | |
| B | 7206 hCoV-19/Wales/PHWC-27D48/2020\|EPI_ISL_432329\|2020- | ···················· | ···················· | ····················· | ···················· | ···················· | ···················· | |
| B | 7207 hCoV-19/Wales/PHWC-2861F/2020\|EPI_ISL_432331\|2020- | ···················· | ···················· | ····················· | ···················· | ···················· | ···················· | |
| B | 7208 hCoV-19/Wales/PHWC-27D1B/2020\|EPI_ISL_432335\|2020- | ···················· | ···················· | ····················· | ···················· | ···················· | ···················· | |
| B | 7209 hCoV-19/Wales/PHWC-24E1B/2020\|EPI_ISL_432336\|2020- | ···················· | ···················· | ····················· | ···················· | ···················· | ···················· | |
| B | 7210 hCoV-19/Wales/PHWC-27B4E/2020\|EPI_ISL_432337\|2020- | ···················· | ···················· | ····················· | ···················· | ···················· | ···················· | |
| B | 7211 hCoV-19/Wales/PHWC-27425/2020\|EPI_ISL_432338\|2020- | ···················· | ···················· | ····················· | ···················· | ···················· | ···················· | |
| B | 7212 hCoV-19/Wales/PHWC-277D1/2020\|EPI_ISL_432339\|2020- | ···················· | ···················· | ····················· | ···················· | ···················· | ···················· | |
| B | 7213 hCoV-19/Wales/PHWC-27461/2020\|EPI_ISL_432340\|2020- | ···················· | ···················· | ····················· | ···················· | ···················· | ···················· | |
| B | 7214 hCoV-19/Wales/PHWC-28451/2020\|EPI_ISL_432342\|2020- | ···················· | ···················· | ····················· | ···················· | ···················· | ···················· | |
| B | 7215 hCoV-19/Wales/PHWC-29FAA/2020\|EPI_ISL_432343\|2020- | ···················· | ···················· | ····················· | ···················· | ···················· | ···················· | |
| B | 7216 hCoV-19/Wales/PHWC-29FF5/2020\|EPI_ISL_432344\|2020- | ···················· | ···················· | ····················· | ···················· | ···················· | ···················· | |
| B | 7217 hCoV-19/Wales/PHWC-2ADFA/2020\|EPI_ISL_432345\|2020- | ···················· | ···················· | ····················· | ···················· | ···················· | ···················· | |
| B | 7218 hCoV-19/Wales/PHWC-27BC6/2020\|EPI_ISL_432346\|2020- | ···················· | ···················· | ····················· | ···················· | ···················· | ···················· | |
| B | 7219 hCoV-19/Wales/PHWC-28460/2020\|EPI_ISL_432347\|2020- | ···················· | ···················· | ····················· | ···················· | ···················· | ···················· | |
| B | 7220 hCoV-19/Wales/PHWC-28891/2020\|EPI_ISL_432349\|2020- | ···················· | ···················· | ····················· | ···················· | ···················· | ···················· | |
| B | 7221 hCoV-19/Wales/PHWC-2799F/2020\|EPI_ISL_432350\|2020- | ···················· | ····A··············· | ····················· | ···················· | ···················· | ···················· | |
| B | 7222 hCoV-19/Wales/PHWC-2866A/2020\|EPI_ISL_432351\|2020- | ···················· | ···················· | ····················· | ···················· | ···················· | ···················· | |
| B | 7223 hCoV-19/Wales/PHWC-283CD/2020\|EPI_ISL_432352\|2020- | ···················· | ···················· | ····················· | ···················· | ···················· | ···················· | |
| B | 7224 hCoV-19/Wales/PHWC-29E25/2020\|EPI_ISL_432353\|2020- | ···················· | ···················· | ····················· | ···················· | ···················· | ···················· | |
| B | 7225 hCoV-19/Wales/PHWC-2AF6D/2020\|EPI_ISL_432354\|2020- | ···················· | ···················· | ····················· | ···················· | ···················· | ···················· | |
| B | 7226 hCoV-19/Wales/PHWC-286D3/2020\|EPI_ISL_432355\|2020- | ···················· | ···················· | ····················· | ···················· | ···················· | ···················· | |
| B | 7227 hCoV-19/Wales/PHWC-286E2/2020\|EPI_ISL_432356\|2020- | ···················· | ···················· | ····················· | ···················· | ···················· | ···················· | |
| B | 7228 hCoV-19/Scotland/CVR1493/2020\|EPI_ISL_433600\|2020- | ···················· | ···················· | ····················· | ···················· | ···················· | ···················· | |
| B | 7229 hCoV-19/Scotland/CVR1498/2020\|EPI_ISL_433602\|2020- | ···················· | ···················· | ····················· | ···················· | ···················· | ···················· | |
| B | 7230 hCoV-19/Scotland/CVR1500/2020\|EPI_ISL_433603\|2020- | ···················· | ···················A | ····················· | ···················· | ···················· | ···················· | 1 |
| B | 7231 hCoV-19/Scotland/CVR1519/2020\|EPI_ISL_433607\|2020- | ···················· | ··············C····· | ····················· | ···················· | ···················· | ···················· | |
| B | 7232 hCoV-19/Scotland/CVR1521/2020\|EPI_ISL_433608\|2020- | ···················· | ···················· | ····················· | ···················· | ···················· | ···················· | |
| B | 7233 hCoV-19/Scotland/CVR1532/2020\|EPI_ISL_433609\|2020- | ···················· | ···················· | ····················· | ···················· | ···················· | ···················· | |
| B | 7234 hCoV-19/Scotland/CVR1550/2020\|EPI_ISL_433614\|2020- | ···················· | ···················· | ····················· | ···················· | ···················· | ···················· | |
| B | 7235 hCoV-19/Scotland/CVR1554/2020\|EPI_ISL_433615\|2020- | ···················· | ···················· | ····················· | ···················· | ···················· | ···················· | |
| B | 7236 hCoV-19/Scotland/CVR1562/2020\|EPI_ISL_433617\|2020- | ···················· | ·R·················R | ····················· | ···················· | ···················· | ···················· | 1 |
| B | 7237 hCoV-19/Scotland/CVR1569/2020\|EPI_ISL_433621\|2020- | ···················· | ···················· | ····················· | ···················· | ···················· | ···················· | |
| B | 7238 hCoV-19/Scotland/CVR1575/2020\|EPI_ISL_433622\|2020- | ···················· | ···················· | ····················· | ···················· | ···················· | ···················· | |
| B | 7239 hCoV-19/Scotland/CVR1588/2020\|EPI_ISL_433626\|2020- | ···················· | ···················A | ····················· | ···················· | ···················· | ···················· | 1 |
| B | 7240 hCoV-19/Scotland/CVR1607/2020\|EPI_ISL_433630\|2020- | ···················· | ·A·················· | ····················· | ···················· | ···················· | ···················· | |
| B | 7241 hCoV-19/Scotland/CVR1622/2020\|EPI_ISL_433632\|2020- | ···················· | ···················A | ····················· | ···················· | ···················· | ···················· | 1 |
| B | 7242 hCoV-19/Scotland/CVR1629/2020\|EPI_ISL_433633\|2020- | ···················· | ·A·················· | ····················· | ···················· | ···················· | ···················· | |
| B | 7243 hCoV-19/Scotland/CVR1633/2020\|EPI_ISL_433636\|2020- | ···················· | ···················· | ····················· | ···················· | ···················· | ···················· | |
| B | 7244 hCoV-19/Scotland/CVR1635/2020\|EPI_ISL_433638\|2020- | ···················· | ·A·················· | ····················· | ···················· | ···················· | ···················· | |
| B | 7245 hCoV-19/Scotland/CVR1639/2020\|EPI_ISL_433640\|2020- | ···················· | ···················· | ····················· | ···················· | ···················· | ···················· | |
| B | 7246 hCoV-19/Scotland/CVR1644/2020\|EPI_ISL_433641\|2020- | ···················· | ···················A | ····················· | ···················· | ···················· | ···················· | 1 |
| B | 7247 hCoV-19/Scotland/CVR1647/2020\|EPI_ISL_433642\|2020- | ···················· | ···················· | ····················· | ···················· | ···················· | ···················· | |
| B | 7248 hCoV-19/Scotland/CVR1652/2020\|EPI_ISL_433643\|2020- | ···················· | ···················· | ····················· | ···················· | ···················· | ···················· | |
| B | 7249 hCoV-19/Scotland/CVR1657/2020\|EPI_ISL_433644\|2020- | ···················· | ···················· | ····················· | ···················· | ···················· | ···················· | |
| B | 7250 hCoV-19/Scotland/CVR1663/2020\|EPI_ISL_433646\|2020- | ···················· | ···················· | ····················· | ···················· | ···················· | ···················· | |
| B | 7251 hCoV-19/Scotland/CVR1669/2020\|EPI_ISL_433647\|2020- | ···················· | ···················· | ····················· | ···················· | ···················· | ···················· | |
| B | 7252 hCoV-19/Scotland/CVR1673/2020\|EPI_ISL_433648\|2020- | ···················· | ···················· | ····················· | ···················· | ···················· | ···················· | |
| B | 7253 hCoV-19/Scotland/CVR1675/2020\|EPI_ISL_433649\|2020- | ···················· | ···················· | ···········T········· | ···················· | ···················· | ···················· | |
| B | 7254 hCoV-19/Scotland/CVR1676/2020\|EPI_ISL_433650\|2020- | ···················· | ···················· | ····················· | ···················· | ···················· | ···················· | |
| B | 7255 hCoV-19/Scotland/CVR1677/2020\|EPI_ISL_433651\|2020- | ···················· | ···················· | ····················· | ···················· | ···················· | ···················· | |
| B | 7256 hCoV-19/Scotland/CVR1685/2020\|EPI_ISL_433653\|2020- | ···················· | ···················A | ····················· | ···················· | ···················· | ···················· | 1 |
| B | 7257 hCoV-19/Scotland/CVR1695/2020\|EPI_ISL_433655\|2020- | ···················· | ···················· | ····················· | ···················· | ···················· | ···················· | |
| B | 7258 hCoV-19/Scotland/CVR1698/2020\|EPI_ISL_433656\|2020- | ···················· | ···················· | ····················· | ···················· | ···················· | ···················· | |
| B | 7259 hCoV-19/Scotland/CVR1710/2020\|EPI_ISL_433657\|2020- | ···················· | ···················· | ····················· | ···················· | ···················· | ···················· | |
| B | 7260 hCoV-19/Scotland/CVR1751/2020\|EPI_ISL_433658\|2020- | ···················· | ···················· | ····················· | ···················· | ···················· | ···················· | |

| | | | | | | | | | |
|---|---|---|---|---|---|---|---|---|---|
| B | 7261 | hCoV-19/Scotland/CVR1935/2020\|EPI_ISL_433663\|2020- | ···················· | ··············C····· | ···················· | ···················· | ···················· | ···················· | |
| B | 7262 | hCoV-19/England/CAMB-73B16/2020\|EPI_ISL_433666\|202 | ···················· | ···················· | ···················· | ···················· | ···················· | ···················· | |
| B | 7263 | hCoV-19/England/CAMB-73B61/2020\|EPI_ISL_433667\|202 | ···················· | ···················· | ···················· | ···················· | ···················· | ···················· | |
| B | 7264 | hCoV-19/England/CAMB-73B70/2020\|EPI_ISL_433668\|202 | ···················· | ···················· | ···················· | ···················· | ···················· | ···················· | |
| B | 7265 | hCoV-19/England/CAMB-73B8F/2020\|EPI_ISL_433669\|202 | ···················· | ···················· | ···················· | ···················· | ···················· | ···················· | |
| B | 7266 | hCoV-19/England/CAMB-73BAD/2020\|EPI_ISL_433671\|202 | ···················· | ···················A | ···················· | ···················· | ···················· | ···················· | 1 |
| B | 7267 | hCoV-19/England/CAMB-73BBC/2020\|EPI_ISL_433672\|202 | ···················· | ···················· | ···················· | ···················· | ···················· | ···················· | |
| B | 7268 | hCoV-19/England/CAMB-73BCB/2020\|EPI_ISL_433673\|202 | ···················· | ···················A | ···················· | ···················· | ···················· | ···················· | 1 |
| B | 7269 | hCoV-19/England/CAMB-73BDA/2020\|EPI_ISL_433674\|202 | ···················· | ···················· | ···················· | ···················· | ···················· | ···················· | |
| B | 7270 | hCoV-19/England/CAMB-73BF8/2020\|EPI_ISL_433675\|202 | ···················· | ···················· | ···················· | ···················· | ···················· | ···················· | |
| B | 7271 | hCoV-19/England/CAMB-73C04/2020\|EPI_ISL_433676\|202 | ···················· | ···················· | ···················· | ···················· | ···················· | ···················· | |
| B | 7272 | hCoV-19/England/CAMB-73C13/2020\|EPI_ISL_433677\|202 | ···················· | ···················· | ···················· | ···················· | ···················· | ···················· | |
| B | 7273 | hCoV-19/England/CAMB-73C31/2020\|EPI_ISL_433678\|202 | ···················· | ···················· | ···················· | ···················· | ···················· | ···················· | |
| B | 7274 | hCoV-19/England/CAMB-73C40/2020\|EPI_ISL_433679\|202 | ···················· | ···················· | ···················· | ···················· | ···················· | ···················· | |
| B | 7275 | hCoV-19/England/CAMB-73C5F/2020\|EPI_ISL_433680\|202 | ···················· | ···················· | ···················· | ···················· | ···················· | ···················· | |
| B | 7276 | hCoV-19/England/CAMB-73C7D/2020\|EPI_ISL_433681\|202 | ···················· | ···················· | ···················· | ···················· | ···················· | ···················· | |
| B | 7277 | hCoV-19/England/CAMB-73C9B/2020\|EPI_ISL_433682\|202 | ···················· | ···················· | ···················· | ···················· | ···················· | ···················· | |
| B | 7278 | hCoV-19/England/CAMB-73CAA/2020\|EPI_ISL_433683\|202 | ···················· | ···················· | ···················· | ···················· | ···················· | ···················· | |
| B | 7279 | hCoV-19/England/CAMB-73CC8/2020\|EPI_ISL_433684\|202 | ···················· | ···················· | ···················· | ···················· | ···················· | ···················· | |
| B | 7280 | hCoV-19/England/CAMB-73CD7/2020\|EPI_ISL_433685\|202 | ···················· | ···················· | ···················· | ···················· | ···················· | ···················· | |
| B | 7281 | hCoV-19/England/CAMB-73D01/2020\|EPI_ISL_433686\|202 | ···················· | ···················· | ···················· | ···················· | ···················· | ···················· | |
| B | 7282 | hCoV-19/England/CAMB-73D10/2020\|EPI_ISL_433687\|202 | ···················· | ···················· | ···················· | ···················· | ···················· | ···················· | |
| B | 7283 | hCoV-19/Belgium/UGent-30/2020\|EPI_ISL_434348\|2020- | ···················· | ···················· | ···················· | ···················· | ···················· | ···················· | |
| B | 7284 | hCoV-19/Belgium/UGent-31/2020\|EPI_ISL_434349\|2020- | ···················· | ···················· | ···················· | ···················· | ···················· | ···················· | |
| B | 7285 | hCoV-19/Belgium/UGent-33/2020\|EPI_ISL_434351\|2020- | ···················· | ···················· | ···················· | ···················· | ···················· | ···················· | |
| B | 7286 | hCoV-19/Belgium/UGent-34/2020\|EPI_ISL_434352\|2020- | ···················· | ···················· | ···················· | ···················· | ···················· | ···················· | |
| B | 7287 | hCoV-19/Belgium/UGent-36/2020\|EPI_ISL_434354\|2020- | ···················· | ···················· | ···················· | ···················· | ···················· | ···················· | |
| B | 7288 | hCoV-19/Belgium/UGent-37/2020\|EPI_ISL_434355\|2020- | ···················· | ···················· | ···················· | ···················· | ···················· | ···················· | |
| B | 7289 | hCoV-19/Belgium/UGent-40/2020\|EPI_ISL_434357\|2020- | ···················· | ···················· | ···········T········ | ···················· | ···················· | ···················· | |
| B | 7290 | hCoV-19/USA/TX-HMH0315/2020\|EPI_ISL_434931\|2020-03 | ···················· | ···················· | ···················· | ···················· | ···················· | ···················· | |
| B | 7291 | hCoV-19/USA/TX-HMH0320/2020\|EPI_ISL_434933\|2020-03 | ···················· | ···················· | ···················· | ···················· | ···················· | ···················· | |
| B | 7292 | hCoV-19/USA/TX-HMH0322/2020\|EPI_ISL_434934\|2020-03 | ···················· | ···················· | ···················· | ···················· | ···················· | ···················· | |
| B | 7293 | hCoV-19/USA/TX-HMH0323/2020\|EPI_ISL_434935\|2020-03 | ···················· | ···················· | ···················· | ···················· | ···················· | ···················· | |
| B | 7294 | hCoV-19/USA/TX-HMH0324/2020\|EPI_ISL_434936\|2020-03 | ···················· | ···················· | ···················· | ···················· | ···················· | ···················· | |
| B | 7295 | hCoV-19/USA/TX-HMH0325/2020\|EPI_ISL_434937\|2020-03 | ···················· | ···················· | ···················· | ···················· | ···················· | ···················· | |
| B | 7296 | hCoV-19/USA/TX-HMH0326/2020\|EPI_ISL_434938\|2020-03 | ···················· | ···················· | ···················· | ···················· | ···················· | ···················· | |
| B | 7297 | hCoV-19/USA/TX-HMH0327/2020\|EPI_ISL_434939\|2020-03 | ···················· | ···················· | ···················· | ···················· | ···················· | ···················· | |
| B | 7298 | hCoV-19/USA/TX-HMH0328/2020\|EPI_ISL_434940\|2020-03 | ···················· | ···················· | ···················· | ···················· | ···················· | ···················· | |
| B | 7299 | hCoV-19/USA/TX-HMH0347/2020\|EPI_ISL_434941\|2020-04 | ···················· | ···················· | ···················· | ···················· | ···················· | ···················· | |
| B | 7300 | hCoV-19/USA/TX-HMH0348/2020\|EPI_ISL_434942\|2020-04 | ···················· | ···················· | ···················· | ···················· | ···················· | ···················· | |
| B | 7301 | hCoV-19/USA/TX-HMH0349/2020\|EPI_ISL_434943\|2020-04 | ···················· | ···················· | ···················· | ···················· | ···················· | ···················· | |
| B | 7302 | hCoV-19/USA/TX-HMH0351/2020\|EPI_ISL_434945\|2020-03 | ···················· | ···················· | ···················· | ···················· | ···················· | ···················· | |
| B | 7303 | hCoV-19/USA/TX-HMH0352/2020\|EPI_ISL_434946\|2020-03 | ···················· | ···················· | ···················· | ···················· | ···················· | ···················· | |
| B | 7304 | hCoV-19/USA/TX-HMH0353/2020\|EPI_ISL_434947\|2020-03 | ···················· | ···················· | ···················· | ···················· | ···················· | ···················· | |
| B | 7305 | hCoV-19/USA/TX-HMH0354/2020\|EPI_ISL_434948\|2020-03 | ···················· | ···················· | ···················· | ···················· | ···················· | ···················· | |
| B | 7306 | hCoV-19/USA/TX-HMH0355/2020\|EPI_ISL_434949\|2020-03 | ···················· | ···················· | ···················· | ···················· | ···················· | ···················· | |
| B | 7307 | hCoV-19/USA/TX-HMH0357/2020\|EPI_ISL_434950\|2020-03 | ···················· | ···················· | ···················· | ···················· | ···················· | ···················· | |
| B | 7308 | hCoV-19/USA/TX-HMH0358/2020\|EPI_ISL_434951\|2020-03 | ···················· | ···················· | ···················· | ···················· | ···················· | ···················· | |
| B | 7309 | hCoV-19/USA/TX-HMH0359/2020\|EPI_ISL_434952\|2020-03 | ···················· | ···················· | ···················· | ···················· | ···················· | ···················· | |
| B | 7310 | hCoV-19/USA/TX-HMH0360/2020\|EPI_ISL_434953\|2020-03 | ···················· | ···················· | ···················· | ···················· | ···················· | ···················· | |
| B | 7311 | hCoV-19/USA/TX-HMH0361/2020\|EPI_ISL_434954\|2020-03 | ···················· | ···················· | ···················· | ···················· | ···················· | ···················· | |
| B | 7312 | hCoV-19/USA/TX-HMH0363/2020\|EPI_ISL_434955\|2020-03 | ···················· | ···················· | ···················· | ···················· | ···················· | ···················· | |
| B | 7313 | hCoV-19/USA/TX-HMH0364/2020\|EPI_ISL_434956\|2020-03 | ···················· | ···················· | ···················· | ···················· | ···················· | ···················· | |
| B | 7314 | hCoV-19/USA/TX-HMH0365/2020\|EPI_ISL_434957\|2020-03 | ···················· | ···················· | ···················· | ···················· | ···················· | ···················· | |
| B | 7315 | hCoV-19/USA/TX-HMH0366/2020\|EPI_ISL_434958\|2020-03 | ···················· | ···················· | ···················· | ···················· | ···················· | ···················· | |
| B | 7316 | hCoV-19/USA/TX-HMH0367/2020\|EPI_ISL_434959\|2020-03 | ···················· | ···················· | ···················· | ···················· | ···················· | ···················· | |
| B | 7317 | hCoV-19/USA/TX-HMH0369/2020\|EPI_ISL_434960\|2020-03 | ···················· | ···················· | ···················· | ···················· | ···················· | ···················· | |
| B | 7318 | hCoV-19/USA/TX-HMH0370/2020\|EPI_ISL_434961\|2020-03 | ···················· | ···················· | ···················· | ···················· | ···················· | ···················· | |
| B | 7319 | hCoV-19/USA/TX-HMH0371/2020\|EPI_ISL_434962\|2020-03 | ···················· | ···················· | ···················· | ···················· | ···················· | ···················· | |
| B | 7320 | hCoV-19/USA/TX-HMH0372/2020\|EPI_ISL_434963\|2020-03 | ···················· | ···················· | ···················· | ···················· | ···················· | ···················· | |
| B | 7321 | hCoV-19/USA/TX-HMH0375/2020\|EPI_ISL_434964\|2020-03 | ···················· | ···················· | ···················· | ···················· | ···················· | ···················· | |
| B | 7322 | hCoV-19/USA/TX-HMH0376/2020\|EPI_ISL_434965\|2020-03 | ···················· | ···················· | ···················· | ···················· | ···T················ | ···················· | |
| B | 7323 | hCoV-19/USA/TX-HMH0378/2020\|EPI_ISL_434966\|2020-04 | ···················· | ···················· | ···················· | ···················· | ···················· | ···················· | |
| B | 7324 | hCoV-19/USA/TX-HMH0379/2020\|EPI_ISL_434967\|2020-03 | ···················· | ···················· | ···················· | ···················· | ···················· | ···················· | |
| B | 7325 | hCoV-19/USA/TX-HMH0381/2020\|EPI_ISL_434969\|2020-03 | ···················· | ···················· | ···················· | ···················· | ···················· | ···················· | |
| B | 7326 | hCoV-19/USA/TX-HMH0382/2020\|EPI_ISL_434970\|2020-04 | ···················· | ···················· | ···················· | ···················· | ···················· | ···················· | |
| B | 7327 | hCoV-19/USA/TX-HMH0383/2020\|EPI_ISL_434971\|2020-03 | ···················· | ···················· | ···················· | ···················· | ···················· | ···················· | |
| B | 7328 | hCoV-19/USA/TX-HMH0384/2020\|EPI_ISL_434972\|2020-03 | ···················· | ···················· | ···················· | ···················· | ···················· | ···················· | |
| B | 7329 | hCoV-19/USA/TX-HMH0385/2020\|EPI_ISL_434973\|2020-03 | ···················· | ···················· | ···················· | ···················· | ···················· | ···················· | |
| B | 7330 | hCoV-19/USA/TX-HMH0387/2020\|EPI_ISL_434974\|2020-03 | ···················· | ···················· | ···················· | ···················· | ···················· | ···················· | |
| B | 7331 | hCoV-19/USA/TX-HMH0388/2020\|EPI_ISL_434975\|2020-03 | ···················· | ···················· | ···················· | ···················· | ···················· | ···················· | |
| B | 7332 | hCoV-19/USA/TX-HMH0389/2020\|EPI_ISL_434976\|2020-04 | ···················· | ···················· | ···················· | ···················· | ···················· | ···················· | |
| B | 7333 | hCoV-19/USA/TX-HMH0390/2020\|EPI_ISL_434977\|2020-03 | ···················· | ···················· | ···················· | ···················· | ···················· | ···················· | |
| B | 7334 | hCoV-19/USA/TX-HMH0391/2020\|EPI_ISL_434978\|2020-03 | ···················· | ···················· | ···················· | ···················· | ···················· | ···················· | |
| B | 7335 | hCoV-19/USA/TX-HMH0392/2020\|EPI_ISL_434979\|2020-03 | ···················· | ···················· | ···················· | ···················· | ···················· | ···················· | |
| B | 7336 | hCoV-19/USA/TX-HMH0393/2020\|EPI_ISL_434980\|2020-03 | ···················· | ···················· | ···················· | ···················· | ···················· | ···················· | |
| B | 7337 | hCoV-19/USA/TX-HMH0394/2020\|EPI_ISL_434981\|2020-03 | ···················· | ···················· | ···················· | ···················· | ···················· | ···················· | |
| B | 7338 | hCoV-19/USA/TX-HMH0395/2020\|EPI_ISL_434982\|2020-04 | ···················· | ···················· | ···················· | ···················· | ···················· | ···················· | |

| | | | | | | | | |
|---|---|---|---|---|---|---|---|---|
| B | 7339 | hCoV-19/USA/TX-HMH0396/2020\|EPI_ISL_434983\|2020-03 | ···················· | ···················· | ···················· | ···················· | ···················· | ···················· |
| B | 7340 | hCoV-19/USA/TX-HMH0397/2020\|EPI_ISL_434984\|2020-03 | ···················· | ···················· | ···················· | ···················· | ···················· | ···················· |
| B | 7341 | hCoV-19/USA/TX-HMH0398/2020\|EPI_ISL_434985\|2020-03 | ···················· | ···················· | ···················· | ···················· | ···················· | ···················· |
| B | 7342 | hCoV-19/USA/TX-HMH0399/2020\|EPI_ISL_434986\|2020-03 | ···················· | ···················· | ···················· | ···················· | ···················· | ···················· |
| B | 7343 | hCoV-19/USA/TX-HMH0400/2020\|EPI_ISL_434987\|2020-04 | ···················· | ···················· | ···················· | ···················· | ···················· | ···················· |
| B | 7344 | hCoV-19/USA/TX-HMH0401/2020\|EPI_ISL_434988\|2020-03 | ···················· | ···················· | ···················· | ···················· | ···················· | ···················· |
| B | 7345 | hCoV-19/USA/TX-HMH0402/2020\|EPI_ISL_434989\|2020-03 | ···················· | ···················· | ···················· | ···················· | ···················· | ···················· |
| B | 7346 | hCoV-19/USA/TX-HMH0408/2020\|EPI_ISL_434994\|2020-03 | ···················· | ···················· | ···················· | ···················· | ···················· | ···················· |
| B | 7347 | hCoV-19/USA/TX-HMH0412/2020\|EPI_ISL_434997\|2020-03 | ···················· | ···················· | ···················· | ···················· | ···················· | ···················· |
| B | 7348 | hCoV-19/USA/TX-HMH0413/2020\|EPI_ISL_434998\|2020-04 | ···················· | ···················· | ···················· | ···················· | ···················· | ···················· |
| B | 7349 | hCoV-19/USA/TX-HMH0415/2020\|EPI_ISL_434999\|2020-04 | ···················· | ···················· | ···················· | ···················· | ···················· | ···················· |
| B | 7350 | hCoV-19/Singapore/74/2020\|EPI_ISL_435679\|2020-04-0 | ···················· | ···················· | ···················· | ···················· | ···················· | ···················· |
| B | 7351 | hCoV-19/Singapore/75/2020\|EPI_ISL_435680\|2020-04-0 | ···················· | ···················· | ···················· | ···················· | ···················· | ···················· |
| B | 7352 | hCoV-19/Singapore/76/2020\|EPI_ISL_435681\|2020-04-0 | ···················· | ···················· | ···················· | ···················· | ···················· | ···················· |
| B | 7353 | hCoV-19/Singapore/77/2020\|EPI_ISL_435682\|2020-04-0 | ···················· | ···················· | ···················· | ···················· | ···················· | ···················· |
| B | 7354 | hCoV-19/Singapore/78/2020\|EPI_ISL_435683\|2020-04-0 | ···················· | ···················· | ···················· | ···················· | ···················· | ···················· |
| B | 7355 | hCoV-19/Singapore/79/2020\|EPI_ISL_435684\|2020-04-1 | ···················· | ···················· | ···················· | ···················· | ···················· | ···················· |
| B | 7356 | hCoV-19/Singapore/80/2020\|EPI_ISL_435685\|2020-04-1 | ···················· | ···················· | ···················· | ···················· | ···················· | ···················· |
| B | 7357 | hCoV-19/Singapore/81/2020\|EPI_ISL_435686\|2020-04-1 | ···················· | ···M················ | ···················· | ···················· | ···················· | ···················· |
| B | 7358 | hCoV-19/Singapore/82/2020\|EPI_ISL_435687\|2020-04-1 | ···················· | ···················· | ···················· | ···················· | ···················· | ···················· |
| B | 7359 | hCoV-19/Singapore/83/2020\|EPI_ISL_435688\|2020-04-1 | ···················· | ···················· | ···················· | ···················· | ···················· | ···················· |
| B | 7360 | hCoV-19/Singapore/84/2020\|EPI_ISL_435689\|2020-04-1 | ···················· | ···················· | ···················· | ···················· | ···················· | ···················· |
| B | 7361 | hCoV-19/England/CAMB-7EFA6/2020\|EPI_ISL_443000\|202 | ···················· | ···················· | ···················· | ···················· | ···················· | ···················· |
| B | 7362 | hCoV-19/England/CAMB-7F497/2020\|EPI_ISL_443001\|202 | ···················· | ···················· | ···················· | ···················· | ···················· | ···················· |
| B | 7363 | hCoV-19/England/CAMB-7EE5E/2020\|EPI_ISL_443002\|202 | ···················· | ···················· | ···················· | ···················· | ···················· | ···················· |
| B | 7364 | hCoV-19/England/CAMB-7EAD2/2020\|EPI_ISL_443003\|202 | ···················· | ···················· | ···················· | ···················· | ···················· | ···················· |
| B | 7365 | hCoV-19/England/CAMB-7EE9A/2020\|EPI_ISL_443004\|202 | ···················· | ···················· | ···················· | ···················· | ···················· | ···················· |
| B | 7366 | hCoV-19/England/BIRM-5E73B/2020\|EPI_ISL_444300\|202 | ···················· | ···················· | ···················· | ···················· | ···················· | ···················· |
| B | 7367 | hCoV-19/England/CAMB-1AAE67/2020\|EPI_ISL_444320\|20 | ···················· | ···················· | ···················· | ···················· | ···················· | ···················· |
| B | 7368 | hCoV-19/England/CAMB-1AB561/2020\|EPI_ISL_444323\|20 | ···················· | ···················· | ···················· | ···················· | ···················· | ···················· |
| B | 7369 | hCoV-19/England/CAMB-1AB701/2020\|EPI_ISL_444328\|20 | ···················· | ···················· | ···················· | ···················· | ···················· | ···················· |
| B | 7370 | hCoV-19/England/CAMB-1AB74D/2020\|EPI_ISL_444329\|20 | ···················· | ···················· | ···················· | ···················· | ···················· | ···················· |
| B | 7371 | hCoV-19/England/CAMB-1AB80E/2020\|EPI_ISL_444334\|20 | ···················· | ···················· | ···················· | ···················· | ···················· | ···················· |
| B | 7372 | hCoV-19/England/CAMB-1AB82C/2020\|EPI_ISL_444335\|20 | ···················· | ···················· | ···················· | ···················· | ···················· | ···················· |
| B | 7373 | hCoV-19/Guam/GU_NHG_03/2020\|EPI_ISL_445000\|2020-03 | ···················· | ···················· | ···················· | ···················· | ···················· | ···················· |
| B | 7374 | hCoV-19/Wales/PHWC-2A677/2020\|EPI_ISL_445601\|2020- | ···················· | ···················· | ···················· | ···················· | ···················· | ···················· |
| B | 7375 | hCoV-19/Wales/PHWC-2A686/2020\|EPI_ISL_445602\|2020- | ···················· | ···················· | ···················· | ···················· | ···················· | ···················· |
| B | 7376 | hCoV-19/Wales/PHWC-2A6C2/2020\|EPI_ISL_445606\|2020- | ···················· | ···················· | ···················· | ···················· | ···················· | ···················· |
| B | 7377 | hCoV-19/Wales/PHWC-2A747/2020\|EPI_ISL_445614\|2020- | ···················· | ···················· | ···················· | ···················· | ···················· | ···················· |
| B | 7378 | hCoV-19/Wales/PHWC-2A792/2020\|EPI_ISL_445617\|2020- | ···················· | ···················· | ···················· | ···················· | ···················· | ···················· |
| B | 7379 | hCoV-19/Wales/PHWC-2A7DE/2020\|EPI_ISL_445619\|2020- | ···················· | ···················· | ···················· | ···················· | ···················· | ···················· |
| B | 7380 | hCoV-19/Wales/PHWC-2A835/2020\|EPI_ISL_445625\|2020- | ···················· | ···················· | ···················· | ···················· | ···················· | ···················· |
| B | 7381 | hCoV-19/Wales/PHWC-2A871/2020\|EPI_ISL_445628\|2020- | ···················· | ···················· | ···················· | ···················· | ···················· | ···················· |
| B | 7382 | hCoV-19/Wales/PHWC-2A8AE/2020\|EPI_ISL_445631\|2020- | ···················· | ···················· | ···················· | ···················· | ···················· | ···················· |
| B | 7383 | hCoV-19/Wales/PHWC-2A8CC/2020\|EPI_ISL_445632\|2020- | ···················· | ···················· | ···················· | ···················· | ···················· | ···················· |
| B | 7384 | hCoV-19/Wales/PHWC-2A950/2020\|EPI_ISL_445640\|2020- | ···················· | ···················· | ···················· | ···················· | ···················· | ···················· |
| B | 7385 | hCoV-19/Wales/PHWC-2A96F/2020\|EPI_ISL_445641\|2020- | ···················· | ···················· | ···················· | ···················· | ···················· | ···················· |
| B | 7386 | hCoV-19/Wales/PHWC-2A97E/2020\|EPI_ISL_445642\|2020- | ···················· | ···················· | ···················· | ···················· | ···················· | ···················· |
| B | 7387 | hCoV-19/Wales/PHWC-2A99C/2020\|EPI_ISL_445644\|2020- | ···················· | ···················· | ···················· | ···················· | ···················· | ···················· |
| B | 7388 | hCoV-19/Wales/PHWC-2A9E7/2020\|EPI_ISL_445649\|2020- | ···················· | ···················· | ···················· | ···················· | ···················· | ···················· |
| B | 7389 | hCoV-19/Wales/PHWC-2A9F6/2020\|EPI_ISL_445650\|2020- | ···················· | ···················· | ···················· | ···················· | ···················· | ···················· |
| B | 7390 | hCoV-19/Wales/PHWC-2AA02/2020\|EPI_ISL_445651\|2020- | ···················· | ···················· | ···················· | ···················· | ···················· | ···················· |
| B | 7391 | hCoV-19/Wales/PHWC-2AA3F/2020\|EPI_ISL_445654\|2020- | ···················· | ···················· | ···················· | ···················· | ···················· | ···················· |
| B | 7392 | hCoV-19/Wales/PHWC-2AAC6/2020\|EPI_ISL_445662\|2020- | ···················· | ···················· | ···················· | ···················· | ···················· | ···················· |
| B | 7393 | hCoV-19/Wales/PHWC-30ABE/2020\|EPI_ISL_446317\|2020- | ···················· | ···················· | ···················· | ···················· | ···················· | ···················· |
| B | 7394 | hCoV-19/Wales/PHWC-30B7F/2020\|EPI_ISL_446328\|2020- | ···················· | ···················· | ···················· | ···················· | ···················· | ···················· |
| B | 7395 | hCoV-19/Wales/PHWC-30BBB/2020\|EPI_ISL_446332\|2020- | ···················· | ···················· | ···················· | ···················· | ···················· | ···················· |
| B | 7396 | hCoV-19/Wales/PHWC-30BE8/2020\|EPI_ISL_446335\|2020- | ···················· | ···················· | ···················· | ···················· | ···················· | ···················· |
| B | 7397 | hCoV-19/Wales/PHWC-30BF7/2020\|EPI_ISL_446336\|2020- | ···················· | ···················· | ···················· | ···················· | ···················· | ···················· |
| B | 7398 | hCoV-19/Wales/PHWC-34425/2020\|EPI_ISL_446920\|2020- | ···················· | ·A·················· | ···················· | ···················· | ···················· | ···················· |
| B | 7399 | hCoV-19/Wales/PHWC-34452/2020\|EPI_ISL_446923\|2020- | ···················· | ···················· | ···················· | ···················· | ···················· | ···················· |
| B | 7400 | hCoV-19/Wales/PHWC-3448F/2020\|EPI_ISL_446926\|2020- | ···················· | ···················· | ···················· | ···················· | ···················· | ···················· |
| B | 7401 | hCoV-19/Wales/PHWC-34504/2020\|EPI_ISL_446934\|2020- | ···················· | ···················· | ···················· | ···················· | ···················· | ···················· |
| B | 7402 | hCoV-19/Wales/PHWC-345AA/2020\|EPI_ISL_446943\|2020- | ···················· | ···················· | ···················· | ···················· | ···················· | ···················· |
| B | 7403 | hCoV-19/Wales/PHWC-34610/2020\|EPI_ISL_446949\|2020- | ···················· | ···················· | ···················· | ···················· | ···················· | ···················· |
| B | 7404 | hCoV-19/Wales/PHWC-34698/2020\|EPI_ISL_446957\|2020- | ···················· | ···················· | ···················· | ···················· | ···················· | ···················· |
| B | 7405 | hCoV-19/Wales/PHWC-346B6/2020\|EPI_ISL_446959\|2020- | ···················· | ···················· | ···················· | ···················· | ···················· | ···················· |
| B | 7406 | hCoV-19/Wales/PHWC-3480B/2020\|EPI_ISL_446979\|2020- | ···················· | ···················· | ···················· | ···················· | ···················· | ···················· |
| B | 7407 | hCoV-19/Wales/PHWC-348A1/2020\|EPI_ISL_446988\|2020- | ···················· | ···················· | ···················· | ···················· | ···················· | ···················· |
| B | 7408 | hCoV-19/Wales/PHWC-348FC/2020\|EPI_ISL_446992\|2020- | ···················· | ···················· | ···················· | ···················· | ···················· | ···················· |
| B | 7409 | hCoV-19/Wales/PHWC-34917/2020\|EPI_ISL_446994\|2020- | ···················· | ···················· | ···················· | ···················· | ···················· | ···················· |
| B | 7410 | hCoV-19/Wales/PHWC-34926/2020\|EPI_ISL_446995\|2020- | ···················· | ···················· | ···················· | ···················· | ···················· | ···················· |
| B | 7411 | hCoV-19/Thailand/Bangkok-0054/2020\|EPI_ISL_446996\| | ···················· | ···················· | ···················· | ···················· | ···················· | ···················· |
| B | 7412 | hCoV-19/Greece/247_32207/2020\|EPI_ISL_447647\|2020- | ···················· | ···················· | ···················· | ···················· | ···················· | ···················· |
| B | 7413 | hCoV-19/Greece/254_32422/2020\|EPI_ISL_447649\|2020- | ···················· | ···················· | ···················· | ···················· | ···················· | ···················· |
| B | 7414 | hCoV-19/France/IDF-40005BH/2020\|EPI_ISL_447658\|202 | ···················· | ···················· | ···················· | ···················· | ···················· | ···················· |
| B | 7415 | hCoV-19/France/IDF-60003NG/2020\|EPI_ISL_447659\|202 | ···················· | ···················· | ···················· | ···················· | ···················· | ···················· |
| B | 7416 | hCoV-19/France/IDF-10036BG/2020\|EPI_ISL_447660\|202 | ···················· | ···················· | ···················· | ···················· | ···················· | ···················· |

| | | | | | | | | |
|---|---|---|---|---|---|---|---|---|
| B | 7417 | hCoV-19/France/IDF-10037CJ/2020\|EPI_ISL_447661\|202 | .................... | .................... | .................... | .................... | .................... | .................... |
| B | 7418 | hCoV-19/France/IDF-40006SG/2020\|EPI_ISL_447662\|202 | .................... | .................... | .................... | .................... | .................... | .................... |
| B | 7419 | hCoV-19/France/IDF-10030NV/2020\|EPI_ISL_447663\|202 | .................... | .................... | .................... | .................... | .................... | .................... |
| B | 7420 | hCoV-19/France/IDF-10040LJ/2020\|EPI_ISL_447664\|202 | .................... | .................... | .................... | .................... | .................... | .................... |
| B | 7421 | hCoV-19/France/IDF-10041MR/2020\|EPI_ISL_447665\|202 | .................... | .................... | .................... | .................... | .................... | .................... |
| B | 7422 | hCoV-19/France/IDF-20001SE/2020\|EPI_ISL_447666\|202 | .................... | .................... | .................... | .................... | .................... | .................... |
| B | 7423 | hCoV-19/France/IDF-10048SA/2020\|EPI_ISL_447667\|202 | .................... | .................... | .................... | .................... | .................... | .................... |
| B | 7424 | hCoV-19/France/IDF-10046GM/2020\|EPI_ISL_447668\|202 | .................... | .................... | .................... | .................... | .................... | .................... |
| B | 7425 | hCoV-19/Wales/PHWC-29CC1/2020\|EPI_ISL_432330\|2020- | .................... | .................... | .................... | .................... | .................... | .................... |
| B | 7426 | hCoV-19/Wales/PHWC-26BE5/2020\|EPI_ISL_432332\|2020- | .................... | .................... | .................... | .................... | .................... | .................... |
| B | 7427 | hCoV-19/Wales/PHWC-2A079/2020\|EPI_ISL_432334\|2020- | .................... | .................... | .................... | .................... | .................... | .................... |
| B | 7428 | hCoV-19/Scotland/CVR1856/2020\|EPI_ISL_433660\|2020- | .................... | .................... | .................... | .................... | .................... | .................... |
| B | 7429 | hCoV-19/Scotland/CVR1961/2020\|EPI_ISL_433664\|2020- | .................... | .................... | .................... | .................... | .................... | .................... |
| B | 7430 | hCoV-19/Scotland/CVR1971/2020\|EPI_ISL_433665\|2020- | .................... | .................... | .................... | .................... | .................... | .................... |
| B | 7431 | hCoV-19/USA/TX-HMH0272/2020\|EPI_ISL_434900\|2020-03 | .................... | .................... | .................... | .................... | .................... | .................... |
| B | 7432 | hCoV-19/USA/TX-HMH0273/2020\|EPI_ISL_434901\|2020-03 | .................... | .................... | .................... | .................... | .................... | .................... |
| B | 7433 | hCoV-19/USA/TX-HMH0274/2020\|EPI_ISL_434902\|2020-03 | .................... | .................... | .................... | .................... | .................... | .................... |
| B | 7434 | hCoV-19/USA/TX-HMH0275/2020\|EPI_ISL_434903\|2020-03 | .................... | .................... | .................... | .................... | .................... | .................... |
| B | 7435 | hCoV-19/USA/TX-HMH0276/2020\|EPI_ISL_434904\|2020-03 | .................... | .................... | .................... | .................... | .................... | .................... |
| B | 7436 | hCoV-19/USA/TX-HMH0277/2020\|EPI_ISL_434905\|2020-03 | .................... | .................... | .................... | .................... | .................... | .................... |
| B | 7437 | hCoV-19/USA/TX-HMH0278/2020\|EPI_ISL_434906\|2020-03 | .................... | .................... | .................... | .................... | .................... | .................... |
| B | 7438 | hCoV-19/USA/TX-HMH0279/2020\|EPI_ISL_434907\|2020-03 | .................... | .................... | .................... | .................... | .................... | .................... |
| B | 7439 | hCoV-19/USA/TX-HMH0280/2020\|EPI_ISL_434908\|2020-03 | .................... | .................... | .................... | .................... | .................... | .................... |
| B | 7440 | hCoV-19/USA/TX-HMH0281/2020\|EPI_ISL_434909\|2020-03 | .................... | .................... | .................... | .................... | .................... | .................... |
| B | 7441 | hCoV-19/USA/TX-HMH0282/2020\|EPI_ISL_434910\|2020-03 | .................... | .................... | .................... | .................... | .................... | .................... |
| B | 7442 | hCoV-19/USA/TX-HMH0283/2020\|EPI_ISL_434911\|2020-03 | .................... | .................... | .................... | .................... | .................... | .................... |
| B | 7443 | hCoV-19/USA/TX-HMH0284/2020\|EPI_ISL_434912\|2020-03 | .................... | .................... | .................... | .................... | .................... | .................... |
| B | 7444 | hCoV-19/USA/TX-HMH0285/2020\|EPI_ISL_434913\|2020-03 | .................... | .................... | .................... | .................... | .................... | .................... |
| B | 7445 | hCoV-19/USA/TX-HMH0287/2020\|EPI_ISL_434914\|2020-03 | .................... | .................... | .................... | .................... | .................... | .................... |
| B | 7446 | hCoV-19/USA/TX-HMH0289/2020\|EPI_ISL_434915\|2020-03 | .................... | .................... | .................... | .................... | .................... | .................... |
| B | 7447 | hCoV-19/USA/TX-HMH0290/2020\|EPI_ISL_434916\|2020-03 | .................... | .................... | .................... | .................... | .................... | .................... |
| B | 7448 | hCoV-19/USA/TX-HMH0291/2020\|EPI_ISL_434917\|2020-03 | .................... | .................... | .................... | .................... | .................... | .................... |
| B | 7449 | hCoV-19/USA/TX-HMH0292/2020\|EPI_ISL_434918\|2020-03 | .................... | .................... | .................... | .................... | .................... | .................... |
| B | 7450 | hCoV-19/USA/TX-HMH0294/2020\|EPI_ISL_434919\|2020-03 | .................... | .................... | .................... | .................... | .................... | .................... |
| B | 7451 | hCoV-19/USA/TX-HMH0295/2020\|EPI_ISL_434920\|2020-03 | .................... | .................... | .................... | .................... | .................... | .................... |
| B | 7452 | hCoV-19/USA/TX-HMH0298/2020\|EPI_ISL_434921\|2020-03 | .................... | .................... | .................... | .................... | .................... | .................... |
| B | 7453 | hCoV-19/USA/TX-HMH0299/2020\|EPI_ISL_434922\|2020-03 | .................... | .................... | .................... | .................... | .................... | .................... |
| B | 7454 | hCoV-19/USA/TX-HMH0300/2020\|EPI_ISL_434923\|2020-03 | .................... | .................... | .................... | .................... | .................... | .................... |
| B | 7455 | hCoV-19/USA/TX-HMH0301/2020\|EPI_ISL_434924\|2020-03 | .................... | .................... | .................... | .................... | .................... | .................... |
| B | 7456 | hCoV-19/USA/TX-HMH0303/2020\|EPI_ISL_434925\|2020-03 | .................... | .................... | .................... | .................... | .................... | .................... |
| B | 7457 | hCoV-19/USA/TX-HMH0305/2020\|EPI_ISL_434926\|2020-03 | .................... | .................... | .................... | .................... | .................... | .................... |
| B | 7458 | hCoV-19/USA/TX-HMH0306/2020\|EPI_ISL_434927\|2020-03 | .................... | .................... | .................... | .................... | .................... | .................... |
| B | 7459 | hCoV-19/USA/TX-HMH0312/2020\|EPI_ISL_434928\|2020-03 | .................... | .................... | .................... | .................... | .................... | .................... |
| B | 7460 | hCoV-19/USA/TX-HMH0314/2020\|EPI_ISL_434930\|2020-03 | .................... | .................... | .................... | .................... | .................... | .................... |
| B | 7461 | hCoV-19/USA/TX-HMH0403/2020\|EPI_ISL_434990\|2020-03 | .................... | .................... | .................... | .................... | .................... | .................... |
| B | 7462 | hCoV-19/USA/TX-HMH0405/2020\|EPI_ISL_434991\|2020-03 | .................... | .................... | .................... | .................... | .................... | .................... |
| B | 7463 | hCoV-19/USA/TX-HMH0406/2020\|EPI_ISL_434992\|2020-03 | .................... | .................... | .................... | .................... | .................... | .................... |
| B | 7464 | hCoV-19/USA/TX-HMH0407/2020\|EPI_ISL_434993\|2020-03 | .................... | .................... | .................... | .................... | .................... | .................... |
| B | 7465 | hCoV-19/USA/TX-HMH0409/2020\|EPI_ISL_434995\|2020-03 | .................... | .................... | .................... | .................... | .................... | .................... |
| B | 7466 | hCoV-19/USA/TX-HMH0410/2020\|EPI_ISL_434996\|2020-03 | .................... | .................... | .................... | .................... | .................... | .................... |
| B | 7467 | hCoV-19/USA/MI-MDHHS-SC20022/2020\|EPI_ISL_437109\|2 | .................... | .................... | .................... | .................... | .................... | .................... |
| B | 7468 | hCoV-19/USA/MI-MDHHS-SC20024/2020\|EPI_ISL_437111\|2 | .................... | .................... | .................... | .................... | .................... | .................... |
| B | 7469 | hCoV-19/USA/MI-MDHHS-SC20026/2020\|EPI_ISL_437113\|2 | .................... | .................... | .................... | .................... | .................... | .................... |
| B | 7470 | hCoV-19/USA/MI-MDHHS-SC20028/2020\|EPI_ISL_437115\|2 | .................... | .................... | .................... | .................... | .................... | .................... |
| B | 7471 | hCoV-19/USA/MI-MDHHS-SC20030/2020\|EPI_ISL_437117\|2 | .................... | .................... | .................... | .................... | .................... | .................... |
| B | 7472 | hCoV-19/USA/MI-MDHHS-SC20033/2020\|EPI_ISL_437120\|2 | .................... | .................... | .................... | .................... | .................... | .................... |
| B | 7473 | hCoV-19/USA/MI-MDHHS-SC20036/2020\|EPI_ISL_437123\|2 | .................... | .................... | .................... | .................... | .................... | .................... |
| B | 7474 | hCoV-19/USA/MI-MDHHS-SC20041/2020\|EPI_ISL_437128\|2 | .................... | .................... | .................... | .................... | .................... | .................... |
| B | 7475 | hCoV-19/USA/MI-MDHHS-SC20044/2020\|EPI_ISL_437131\|2 | .................... | .................... | .................... | .................... | .................... | .................... |
| B | 7476 | hCoV-19/USA/MI-MDHHS-SC20045/2020\|EPI_ISL_437132\|2 | .................... | .................... | .................... | .................... | .................... | .................... |
| B | 7477 | hCoV-19/USA/MI-MDHHS-SC20048/2020\|EPI_ISL_437135\|2 | .................... | .................... | .................... | .................... | .................... | .................... |
| B | 7478 | hCoV-19/USA/MI-MDHHS-SC20050/2020\|EPI_ISL_437137\|2 | .................... | .................... | .................... | .................... | .................... | .................... |
| B | 7479 | hCoV-19/USA/MI-MDHHS-SC20051/2020\|EPI_ISL_437138\|2 | .................... | .................... | .................... | .................... | .................... | .................... |
| B | 7480 | hCoV-19/USA/MI-MDHHS-SC20052/2020\|EPI_ISL_437139\|2 | .................... | .................... | .................... | .................... | .................... | .................... |
| B | 7481 | hCoV-19/USA/MI-MDHHS-SC20053/2020\|EPI_ISL_437140\|2 | .................... | .................... | .................... | .................... | .................... | .................... |
| B | 7482 | hCoV-19/USA/MI-MDHHS-SC20055/2020\|EPI_ISL_437142\|2 | .................... | .................... | .................... | .................... | .................... | .................... |
| B | 7483 | hCoV-19/USA/MI-MDHHS-SC20058/2020\|EPI_ISL_437145\|2 | .................... | .................... | .................... | .................... | .................... | .................... |
| B | 7484 | hCoV-19/USA/MI-MDHHS-SC20059/2020\|EPI_ISL_437146\|2 | .................... | .................... | .................... | .................... | .................... | .................... |
| B | 7485 | hCoV-19/USA/MI-MDHHS-SC20060/2020\|EPI_ISL_437147\|2 | .................... | .................... | .................... | .................... | .................... | .................... |
| B | 7486 | hCoV-19/USA/MI-MDHHS-SC20063/2020\|EPI_ISL_437150\|2 | .................... | .................... | .................... | .................... | .................... | .................... |
| B | 7487 | hCoV-19/USA/MI-MDHHS-SC20065/2020\|EPI_ISL_437152\|2 | .................... | .................... | .................... | .................... | .................... | .................... |
| B | 7488 | hCoV-19/USA/MI-MDHHS-SC20066/2020\|EPI_ISL_437153\|2 | .................... | .................... | .................... | .................... | .................... | .................... |
| B | 7489 | hCoV-19/USA/MI-MDHHS-SC20067/2020\|EPI_ISL_437154\|2 | .................... | .................... | .................... | .................... | .................... | .................... |
| B | 7490 | hCoV-19/USA/MI-MDHHS-SC20068/2020\|EPI_ISL_437155\|2 | .................... | .................... | .................... | .................... | .................... | .................... |
| B | 7491 | hCoV-19/USA/MI-MDHHS-SC20070/2020\|EPI_ISL_437157\|2 | .................... | .................... | .................... | .................... | .................... | .................... |
| B | 7492 | hCoV-19/USA/MI-MDHHS-SC20075/2020\|EPI_ISL_437162\|2 | .................... | .................... | .................... | .................... | .................... | .................... |
| B | 7493 | hCoV-19/USA/MI-MDHHS-SC20076/2020\|EPI_ISL_437163\|2 | .................... | .................... | .................... | .................... | .................... | .................... |
| B | 7494 | hCoV-19/USA/MI-MDHHS-SC20077/2020\|EPI_ISL_437164\|2 | .................... | .................... | .................... | .................... | .................... | .................... |

B 7495 hCoV-19/USA/MI-MDHHS-SC20079/2020|EPI_ISL_437166|2 .................... .................... ..................... .................... .................... ....................
B 7496 hCoV-19/USA/MI-MDHHS-SC20083/2020|EPI_ISL_437170|2 .................... .................... ..................... .................... .................... ....................
B 7497 hCoV-19/USA/MI-MDHHS-SC20087/2020|EPI_ISL_437174|2 .................... .................... ..................... .................... .................... ....................
B 7498 hCoV-19/USA/MI-MDHHS-SC20089/2020|EPI_ISL_437176|2 .................... .................... ..................... .................... .................... ....................
B 7499 hCoV-19/USA/MI-MDHHS-SC20093/2020|EPI_ISL_437180|2 .................... .................... ..................... .................... .................... ....................
B 7500 hCoV-19/Indonesia/JI-ITD-853Sp/2020|EPI_ISL_437187 .................... .................... ..................... .................... .................... ....................
B 7501 hCoV-19/Indonesia/JI-ITD-3590NT/2020|EPI_ISL_43718 .................... .................... ..................... .................... .................... ....................
B 7502 hCoV-19/Indonesia/JK-EIJK-01/2020|EPI_ISL_437189|2 .................... .................... ..................... .................... .................... ....................
B 7503 hCoV-19/Indonesia/JK-EIJK-02/2020|EPI_ISL_437190|2 .................... .................... ..................... .................... .................... ....................
B 7504 hCoV-19/Indonesia/JK-EIJK-03/2020|EPI_ISL_437191|2 .................... .................... ..................... .................... .................... ....................
B 7505 hCoV-19/Indonesia/JK-EIJK-04/2020|EPI_ISL_437192|2 .................... .................... ..................... .................... .................... ....................
B 7506 hCoV-19/DRC/2363/2020|EPI_ISL_437194|2020-04-14 .................... .................... ..................... .................... .................... ....................
B 7507 hCoV-19/Austria/Graz-MUG1/2020|EPI_ISL_437197|2020 .................... .................... ..................... .................... .................... ....................
B 7508 hCoV-19/Austria/Graz-MUG2/2020|EPI_ISL_437198|2020 .................... .................... ..................... .................... .................... ....................
B 7509 hCoV-19/Austria/Graz-MUG3/2020|EPI_ISL_437199|2020 .................... .................... ..................... .................... .................... ....................
B 7510 hCoV-19/England/CAMB-763A2/2020|EPI_ISL_438400|202 .................... .................... ..................... .................... .................... ....................
B 7511 hCoV-19/England/CAMB-763FD/2020|EPI_ISL_438401|202 .................... .................... ..................... .................... .................... ....................
B 7512 hCoV-19/England/CAMB-76366/2020|EPI_ISL_438403|202 .................... .................... ..................... .................... .................... ....................
B 7513 hCoV-19/England/CAMB-71DC7/2020|EPI_ISL_438404|202 .................... .................... ..................... .................... .................... ....................
B 7514 hCoV-19/England/CAMB-75A17/2020|EPI_ISL_438406|202 .................... .................... ..................... .................... .................... ....................
B 7515 hCoV-19/England/CAMB-76C2F/2020|EPI_ISL_438407|202 .................... .................... ..................... .................... .................... ....................
B 7516 hCoV-19/England/CAMB-76524/2020|EPI_ISL_438408|202 .................... .................... ..................... .................... .................... ....................
B 7517 hCoV-19/England/CAMB-76CB6/2020|EPI_ISL_438409|202 .................... .................... ..................... .................... .................... ....................
B 7518 hCoV-19/England/CAMB-761E4/2020|EPI_ISL_438411|202 .................... .............A...... ..................... .................... .................... ....................
B 7519 hCoV-19/England/CAMB-7620F/2020|EPI_ISL_438412|202 .................... .................... ..................... .................... .................... ....................
B 7520 hCoV-19/England/CAMB-762E1/2020|EPI_ISL_438413|202 .................... .................... ..................... .................... .................... ....................
B 7521 hCoV-19/England/CAMB-75FBE/2020|EPI_ISL_438414|202 .................... .................... ..................... .................... .................... ....................
B 7522 hCoV-19/England/CAMB-76005/2020|EPI_ISL_438415|202 .................... .................... ..................... .................... .................... ....................
B 7523 hCoV-19/England/CAMB-75D0F/2020|EPI_ISL_438416|202 .................... .................... ..................... .................... .................... ....................
B 7524 hCoV-19/England/CAMB-75938/2020|EPI_ISL_438419|202 .................... .............A...... ..................... .................... .................... ....................
B 7525 hCoV-19/England/CAMB-76C98/2020|EPI_ISL_438426|202 .................... .................... ..................... .................... .................... ....................
B 7526 hCoV-19/England/CAMB-767C4/2020|EPI_ISL_438427|202 .................... .................... ..................... .................... .................... ....................
B 7527 hCoV-19/England/CAMB-76A70/2020|EPI_ISL_438428|202 .................... .................... ..................... .................... .................... ....................
B 7528 hCoV-19/England/CAMB-76409/2020|EPI_ISL_438440|202 .................... .................... ..................... .................... .................... ....................
B 7529 hCoV-19/England/CAMB-7624B/2020|EPI_ISL_438441|202 .................... .................... ..................... .................... .................... ....................
B 7530 hCoV-19/England/CAMB-76023/2020|EPI_ISL_438442|202 .................... .................... ..................... .................... .................... ....................
B 7531 hCoV-19/England/CAMB-768C1/2020|EPI_ISL_438443|202 .................... .................... ..................... .................... .................... ....................
B 7532 hCoV-19/England/CAMB-76779/2020|EPI_ISL_438446|202 .................... .................... ..................... .................... .................... ....................
B 7533 hCoV-19/England/CAMB-765E8/2020|EPI_ISL_438447|202 .................... .................... ..................... .................... .................... ....................
B 7534 hCoV-19/England/CAMB-75ADB/2020|EPI_ISL_438448|202 .................... .................... ..................... .................... .................... ....................
B 7535 hCoV-19/England/CAMB-75CF3/2020|EPI_ISL_438451|202 .................... .................... ..................... .................... .................... ....................
B 7536 hCoV-19/England/CAMB-761D5/2020|EPI_ISL_438452|202 .................... .................... ..................... .................... .................... ....................
B 7537 hCoV-19/England/CAMB-764FA/2020|EPI_ISL_438453|202 .................... .................... ..................... .................... .................... ....................
B 7538 hCoV-19/England/CAMB-76A16/2020|EPI_ISL_438454|202 .................... .................... ..................... .................... .................... ....................
B 7539 hCoV-19/England/CAMB-75E1B/2020|EPI_ISL_438455|202 .................... .A.................. ..................... .................... .................... ....................
B 7540 hCoV-19/England/CAMB-76A25/2020|EPI_ISL_438456|202 .................... .................... ..................... .................... .................... ....................
B 7541 hCoV-19/England/CAMB-7680D/2020|EPI_ISL_438457|202 .................... .................... ..................... .................... .................... ....................
B 7542 hCoV-19/England/CAMB-71DB8/2020|EPI_ISL_438458|202 .................... .................... ..................... .................... .................... ....................
B 7543 hCoV-19/England/CAMB-76C7A/2020|EPI_ISL_438459|202 .................... .................... ..................... .................... .................... ....................
B 7544 hCoV-19/England/CAMB-76427/2020|EPI_ISL_438460|202 .................... .................... ..................... .................... .................... ....................
B 7545 hCoV-19/England/CAMB-768FE/2020|EPI_ISL_438462|202 .................... .................... ..................... .................... .................... ....................
B 7546 hCoV-19/England/CAMB-75C6C/2020|EPI_ISL_438463|202 .................... .................... ..................... .................... .................... ....................
B 7547 hCoV-19/England/CAMB-7617B/2020|EPI_ISL_438464|202 .................... .................... ..................... .................... .................... ....................
B 7548 hCoV-19/England/CAMB-76AE9/2020|EPI_ISL_438465|202 .................... .................... ..................... .................... .................... ....................
B 7549 hCoV-19/England/CAMB-766C7/2020|EPI_ISL_438466|202 .................... .................... ..................... .................... .................... ....................
B 7550 hCoV-19/England/CAMB-75A26/2020|EPI_ISL_438467|202 .................... .................... ..................... .................... .................... ....................
B 7551 hCoV-19/England/CAMB-76858/2020|EPI_ISL_438468|202 .................... .................... ..................... .................... .................... ....................
B 7552 hCoV-19/England/CAMB-76876/2020|EPI_ISL_438469|202 .................... .................... ..................... .................... .................... ....................
B 7553 hCoV-19/England/CAMB-75983/2020|EPI_ISL_438470|202 .................... .................... ..................... .................... .................... ....................
B 7554 hCoV-19/England/CAMB-75EB1/2020|EPI_ISL_438471|202 .................... .................... ..................... .................... .................... ....................
B 7555 hCoV-19/England/CAMB-75B6F/2020|EPI_ISL_438472|202 .................... .................... ..................... .................... .................... ....................
B 7556 hCoV-19/England/CAMB-75A08/2020|EPI_ISL_438474|202 .................... .................... ..................... .................... .................... ....................
B 7557 hCoV-19/England/CAMB-76515/2020|EPI_ISL_438475|202 .................... .................... ..................... .................... .................... ....................
B 7558 hCoV-19/England/CAMB-759ED/2020|EPI_ISL_438476|202 .................... .................... ..................... .................... .................... ....................
B 7559 hCoV-19/England/CAMB-75EFD/2020|EPI_ISL_438477|202 .................... .................... ..................... .................... .................... ....................
B 7560 hCoV-19/England/CAMB-76D0E/2020|EPI_ISL_438479|202 .................... .................... ..................... .................... .................... ....................
B 7561 hCoV-19/England/CAMB-75FDC/2020|EPI_ISL_438480|202 .................... .................... ..................... .................... .................... ....................
B 7562 hCoV-19/England/CAMB-75CC6/2020|EPI_ISL_438481|202 .................... .................... ..................... .................... .................... ....................
B 7563 hCoV-19/England/CAMB-75947/2020|EPI_ISL_438482|202 .................... .................... ..................... .................... .................... ....................
B 7564 hCoV-19/England/CAMB-76BC8/2020|EPI_ISL_438484|202 .................... .................... ..................... .................... .................... ....................
B 7565 hCoV-19/England/CAMB-76A9E/2020|EPI_ISL_438485|202 .................... .............A...... ..................... .................... .................... ....................
B 7566 hCoV-19/England/CAMB-76C01/2020|EPI_ISL_438486|202 .................... .................... ..................... .................... .................... ....................
B 7567 hCoV-19/England/CAMB-769A0/2020|EPI_ISL_438488|202 .................... .................... ..................... .................... .................... ....................
B 7568 hCoV-19/England/CAMB-759DE/2020|EPI_ISL_438489|202 .................... .................... ..................... .................... .................... ....................
B 7569 hCoV-19/England/CAMB-75DF0/2020|EPI_ISL_438491|202 .................... .................... ..................... .................... .................... ....................
B 7570 hCoV-19/England/CAMB-762C3/2020|EPI_ISL_438492|202 .................... .................... ..................... .................... .................... ....................
B 7571 hCoV-19/England/CAMB-765F7/2020|EPI_ISL_438493|202 .................... .................... ..................... .................... .................... ....................
B 7572 hCoV-19/England/CAMB-767F1/2020|EPI_ISL_438496|202 .................... .................... ..................... .................... .................... ....................

| | | | | | | | |
|---|---|---|---|---|---|---|---|
| B | 7573 hCoV-19/England/CAMB-75E57/2020\|EPI_ISL_438497\|202 | ···················· | ···················· | ···················· | ···················· | ···················· | ···················· |
| B | 7574 hCoV-19/England/CAMB-75DB4/2020\|EPI_ISL_438498\|202 | ···················· | ···················· | ···················· | ···················· | ···················· | ···················· |
| B | 7575 hCoV-19/England/CAMB-76BB9/2020\|EPI_ISL_438499\|202 | ···················· | ···················· | ···················· | ···················· | ···················· | ···················· |
| B | 7576 hCoV-19/England/LIVE-9929D/2020\|EPI_ISL_439706\|202 | ···················· | ···················· | ···················· | ···················· | ···················· | ···················· |
| B | 7577 hCoV-19/England/LIVE-992AC/2020\|EPI_ISL_439707\|202 | ···················· | ···················· | ···················· | ···················· | ···················· | ···················· |
| B | 7578 hCoV-19/England/LIVE-992BB/2020\|EPI_ISL_439708\|202 | ···················· | ···················· | ···················· | ···················· | ···················· | ···················· |
| B | 7579 hCoV-19/England/LIVE-992CA/2020\|EPI_ISL_439709\|202 | ···················· | ···················· | ···················· | ···················· | ···················· | ···················· |
| B | 7580 hCoV-19/England/LIVE-992D9/2020\|EPI_ISL_439710\|202 | ···················· | ···················· | ···················· | ···················· | ···················· | ···················· |
| B | 7581 hCoV-19/England/LIVE-992F7/2020\|EPI_ISL_439712\|202 | ···················· | ···················· | ···················· | ···················· | ···················· | ···················· |
| B | 7582 hCoV-19/England/LIVE-9939A/2020\|EPI_ISL_439714\|202 | ···················· | ···················· | ···················· | ···················· | ···················· | ···················· |
| B | 7583 hCoV-19/England/LIVE-993B8/2020\|EPI_ISL_439715\|202 | ···················· | ···················· | ···················· | ···················· | ···················· | ···················· |
| B | 7584 hCoV-19/England/LIVE-993E5/2020\|EPI_ISL_439718\|202 | ···················· | ···················· | ···················· | ···················· | ···················· | ···················· |
| B | 7585 hCoV-19/England/LIVE-993F4/2020\|EPI_ISL_439719\|202 | ···················· | ···················· | ···················· | ···················· | ···················· | ···················· |
| B | 7586 hCoV-19/England/LIVE-99400/2020\|EPI_ISL_439720\|202 | ···················· | ···················· | ···················· | ···················· | ···················· | ···················· |
| B | 7587 hCoV-19/England/LIVE-9941F/2020\|EPI_ISL_439721\|202 | ···················· | ···················· | ···················· | ···················· | ···················· | ···················· |
| B | 7588 hCoV-19/England/LIVE-9942E/2020\|EPI_ISL_439722\|202 | ···················· | ···················· | ···················· | ···················· | ···················· | ···················· |
| B | 7589 hCoV-19/England/LIVE-9944C/2020\|EPI_ISL_439724\|202 | ···················· | ···················· | ···················· | ···················· | ···················· | ···················· |
| B | 7590 hCoV-19/England/LIVE-9945B/2020\|EPI_ISL_439725\|202 | ···················· | ···················· | ···················· | ···················· | ···················· | ···················· |
| B | 7591 hCoV-19/England/LIVE-994B5/2020\|EPI_ISL_439728\|202 | ···················· | ···················· | ···················· | ···················· | ···················· | ···················· |
| B | 7592 hCoV-19/England/LIVE-994F1/2020\|EPI_ISL_439730\|202 | ···················· | ···················· | ···················· | ···················· | ···················· | ···················· |
| B | 7593 hCoV-19/England/LIVE-9951C/2020\|EPI_ISL_439731\|202 | ···················· | ···················· | ···················· | ···················· | ···················· | ···················· |
| B | 7594 hCoV-19/England/LIVE-9953A/2020\|EPI_ISL_439733\|202 | ···················· | ···················· | ···················· | ···················· | ···················· | ···················· |
| B | 7595 hCoV-19/England/LIVE-995B2/2020\|EPI_ISL_439737\|202 | ···················· | ···················· | ···················· | ···················· | ···················· | ···················· |
| B | 7596 hCoV-19/England/LIVE-99646/2020\|EPI_ISL_439744\|202 | ···················· | ·A·················· | ···················· | ···················· | ···················· | ···················· |
| B | 7597 hCoV-19/England/LIVE-99691/2020\|EPI_ISL_439747\|202 | ···················· | ···················· | ···················· | ···················· | ···················· | ···················· |
| B | 7598 hCoV-19/England/LIVE-996A0/2020\|EPI_ISL_439748\|202 | ···················· | ···················· | ···················· | ···················· | ···················· | ···················· |
| B | 7599 hCoV-19/England/LIVE-9979E/2020\|EPI_ISL_439758\|202 | ···················· | ···················· | ···················· | ···················· | ···················· | ···················· |
| B | 7600 hCoV-19/England/LIVE-998C8/2020\|EPI_ISL_439770\|202 | ···················· | ···················· | ···················· | ···················· | ···················· | ···················· |
| B | 7601 hCoV-19/England/LIVE-9993E/2020\|EPI_ISL_439775\|202 | ···················· | ···················· | ···················· | ···················· | ···················· | ···················· |
| B | 7602 hCoV-19/England/LIVE-9995C/2020\|EPI_ISL_439777\|202 | ···················· | ···················· | ···················· | ···················· | ···················· | ···················· |
| B | 7603 hCoV-19/England/LIVE-99998/2020\|EPI_ISL_439780\|202 | ···················· | ···················· | ···················· | ···················· | ···················· | ···················· |
| B | 7604 hCoV-19/England/LIVE-999B6/2020\|EPI_ISL_439782\|202 | ···················· | ···················· | ···················· | ···················· | ···················· | ···················· |
| B | 7605 hCoV-19/England/LIVE-999D4/2020\|EPI_ISL_439784\|202 | ···················· | ···················· | ···················· | ···················· | ···················· | ···················· |
| B | 7606 hCoV-19/England/LIVE-999E3/2020\|EPI_ISL_439785\|202 | ············T······· | ···················· | ···················· | ···················· | ···················· | ···················· |
| B | 7607 hCoV-19/England/LIVE-99A1D/2020\|EPI_ISL_439787\|202 | ···················· | ···················· | ···················· | ···················· | ···················· | ···················· |
| B | 7608 hCoV-19/England/LIVE-99A2C/2020\|EPI_ISL_439788\|202 | ···················· | ···················· | ···················· | ···················· | ···················· | ···················· |
| B | 7609 hCoV-19/England/LIVE-99A68/2020\|EPI_ISL_439790\|202 | ···················· | ···················· | ···················· | ···················· | ···················· | ···················· |
| B | 7610 hCoV-19/England/LIVE-99A77/2020\|EPI_ISL_439791\|202 | ···················· | ···················· | ···················· | ···················· | ···················· | ···················· |
| B | 7611 hCoV-19/England/LIVE-99AA4/2020\|EPI_ISL_439793\|202 | ···················· | ···················· | ···················· | ···················· | ···················· | ···················· |
| B | 7612 hCoV-19/England/LIVE-99AC2/2020\|EPI_ISL_439795\|202 | ···················· | ·A·················· | ···················· | ···················· | ···················· | ···················· |
| B | 7613 hCoV-19/England/LIVE-99AD1/2020\|EPI_ISL_439796\|202 | ···················· | ···················· | ···················· | ···················· | ···················· | ···················· |
| B | 7614 hCoV-19/England/LIVE-99AFF/2020\|EPI_ISL_439798\|202 | ···················· | ···················· | ···················· | ···················· | ···················· | ···················· |
| B | 7615 hCoV-19/England/LIVE-99B29/2020\|EPI_ISL_439799\|202 | ···················· | ···················· | ···················· | ···················· | ···················· | ···················· |
| B | 7616 hCoV-19/England/BRIS-122FEC/2020\|EPI_ISL_440700\|20 | ···················· | ···················· | ···················· | ···················· | ···················· | ···················· |
| B | 7617 hCoV-19/England/BRIS-122E94/2020\|EPI_ISL_440701\|20 | ···················· | ···················· | ···················· | ···················· | ···················· | ···················· |
| B | 7618 hCoV-19/England/BRIS-122D79/2020\|EPI_ISL_440702\|20 | ···················· | ···················· | ···················· | ···················· | ···················· | ···················· |
| B | 7619 hCoV-19/England/BRIS-122CA9/2020\|EPI_ISL_440703\|20 | ···················· | ···················· | ···················· | ···················· | ···················· | ···················· |
| B | 7620 hCoV-19/England/BRIS-12284B/2020\|EPI_ISL_440704\|20 | ···················· | ···················· | ···················· | ···················· | ···················· | ···················· |
| B | 7621 hCoV-19/England/BRIS-122B9D/2020\|EPI_ISL_440705\|20 | ···················· | ···················· | ···················· | ···················· | ···················· | ···················· |
| B | 7622 hCoV-19/England/BRIS-1229A2/2020\|EPI_ISL_440707\|20 | ···················· | ···················· | ···················· | ···················· | ···················· | ···················· |
| B | 7623 hCoV-19/England/BRIS-122E1C/2020\|EPI_ISL_440710\|20 | ···················· | ·············A······ | ···················· | ···················· | ···················· | ···················· |
| B | 7624 hCoV-19/England/BRIS-12280F/2020\|EPI_ISL_440711\|20 | ···················· | ···················· | ···················· | ···················· | ···················· | ···················· |
| B | 7625 hCoV-19/England/BRIS-122887/2020\|EPI_ISL_440712\|20 | ···················· | ···················· | ···················· | ···················· | ···················· | ···················· |
| B | 7626 hCoV-19/England/BRIS-122C30/2020\|EPI_ISL_440715\|20 | ···················· | ···················· | ···················· | ···················· | ···················· | ···················· |
| B | 7627 hCoV-19/England/BRIS-122F91/2020\|EPI_ISL_440716\|20 | ···················· | ···················· | ···················· | ···················· | ···················· | ···················· |
| B | 7628 hCoV-19/England/BRIS-12278A/2020\|EPI_ISL_440717\|20 | ···················· | ···················· | ···················· | ···················· | ···················· | ···················· |
| B | 7629 hCoV-19/England/BRIS-122447/2020\|EPI_ISL_440718\|20 | ···················· | ···················· | ···················· | ···················· | ···················· | ···················· |
| B | 7630 hCoV-19/England/BRIS-12259F/2020\|EPI_ISL_440719\|20 | ···················· | ···················· | ···················· | ···················· | ···················· | ···················· |
| B | 7631 hCoV-19/England/BRIS-12275D/2020\|EPI_ISL_440721\|20 | ···················· | ···················· | ···················· | ···················· | ···················· | ···················· |
| B | 7632 hCoV-19/England/BRIS-1228D2/2020\|EPI_ISL_440723\|20 | ···················· | ···················· | ···················· | ···················· | ···················· | ···················· |
| B | 7633 hCoV-19/England/BRIS-1228B4/2020\|EPI_ISL_440724\|20 | ···················· | ···················· | ···················· | ···················· | ···················· | ···················· |
| B | 7634 hCoV-19/England/BRIS-122DD3/2020\|EPI_ISL_440726\|20 | ···················· | ···················· | ···················· | ···················· | ···················· | ···················· |
| B | 7635 hCoV-19/England/BRIS-1226E7/2020\|EPI_ISL_440728\|20 | ···················· | ·A·················· | ···················· | ···················· | ···················· | ···················· |
| B | 7636 hCoV-19/England/BRIS-122939/2020\|EPI_ISL_440729\|20 | ···················· | ···················· | ···················· | ···················· | ···················· | ···················· |
| B | 7637 hCoV-19/England/BRIS-122FFB/2020\|EPI_ISL_440736\|20 | ···················· | ···················· | ···················· | ···················· | ···················· | ···················· |
| B | 7638 hCoV-19/England/BRIS-12276C/2020\|EPI_ISL_440737\|20 | ···················· | ···················· | ···················· | ···················· | ···················· | ···················· |
| B | 7639 hCoV-19/England/BRIS-122BCA/2020\|EPI_ISL_440739\|20 | ···················· | ···················· | ···················· | ···················· | ···················· | ···················· |
| B | 7640 hCoV-19/England/BRIS-122EC1/2020\|EPI_ISL_440751\|20 | ···················· | ···················· | ···················· | ···················· | ···················· | ···················· |
| B | 7641 hCoV-19/England/BRIS-122A54/2020\|EPI_ISL_440752\|20 | ···················· | ···················· | ···················· | ···················· | ···················· | ···················· |
| B | 7642 hCoV-19/England/BRIS-122BE8/2020\|EPI_ISL_440753\|20 | ···················· | ···················· | ···················· | ···················· | ···················· | ···················· |
| B | 7643 hCoV-19/England/BRIS-12241A/2020\|EPI_ISL_440754\|20 | ···················· | ···················· | ···················· | ···················· | ···················· | ···················· |
| B | 7644 hCoV-19/England/BRIS-122E76/2020\|EPI_ISL_440755\|20 | ···················· | ···················· | ···················· | ···················· | ···················· | ···················· |
| B | 7645 hCoV-19/England/BRIS-122474/2020\|EPI_ISL_440757\|20 | ···················· | ···················· | ···················· | ···················· | ···················· | ···················· |
| B | 7646 hCoV-19/England/BRIS-1223E0/2020\|EPI_ISL_440758\|20 | ···················· | ···················· | ···················· | ···················· | ···················· | ···················· |
| B | 7647 hCoV-19/England/BRIS-122EFE/2020\|EPI_ISL_440759\|20 | ···················· | ···················· | ···················· | ···················· | ···················· | ···················· |
| B | 7648 hCoV-19/England/BRIS-122544/2020\|EPI_ISL_440761\|20 | ···················· | ···················· | ···················· | ···················· | ···················· | ···················· |
| B | 7649 hCoV-19/England/BRIS-122BD9/2020\|EPI_ISL_440762\|20 | ···················· | ···················· | ···················· | ···················· | ···················· | ···················· |
| B | 7650 hCoV-19/England/BRIS-122A27/2020\|EPI_ISL_440763\|20 | ···················· | ···················· | ···················· | ···················· | ···················· | ···················· |

| | | | | | | | | |
|---|---|---|---|---|---|---|---|---|
| B | 7651 | hCoV-19/England/BRIS-1228A5/2020\|EPI_ISL_440764\|20 | ···················· | ···················· | ····················· | ···················· | ···················· | ···················· |
| B | 7652 | hCoV-19/England/BRIS-122C12/2020\|EPI_ISL_440765\|20 | ···················· | ···················· | ····················· | ···················· | ···················· | ···················· |
| B | 7653 | hCoV-19/England/BRIS-122F28/2020\|EPI_ISL_440766\|20 | ···················· | ···················· | ····················· | ···················· | ···················· | ···················· |
| B | 7654 | hCoV-19/England/BRIS-122957/2020\|EPI_ISL_440767\|20 | ···················· | ···················· | ····················· | ···················· | ···················· | ···················· |
| B | 7655 | hCoV-19/England/BRIS-1225EA/2020\|EPI_ISL_440768\|20 | ···················· | ···················· | ····················· | ···················· | ···················· | ···················· |
| B | 7656 | hCoV-19/England/BRIS-122B60/2020\|EPI_ISL_440770\|20 | ···················· | ···················· | ····················· | ···················· | ···················· | ···················· |
| B | 7657 | hCoV-19/England/BRIS-122465/2020\|EPI_ISL_440773\|20 | ···················· | ···················· | ····················· | ···················· | ···················· | ···················· |
| B | 7658 | hCoV-19/England/BRIS-1229B1/2020\|EPI_ISL_440774\|20 | ···················· | ···················· | ····················· | ···················· | ···················· | ···················· |
| B | 7659 | hCoV-19/England/BRIS-122CC7/2020\|EPI_ISL_440775\|20 | ···················· | ···················· | ····················· | ···················· | ···················· | ···················· |
| B | 7660 | hCoV-19/England/BRIS-1227A8/2020\|EPI_ISL_440776\|20 | ···················· | ···················· | ····················· | ···················· | ···················· | ···················· |
| B | 7661 | hCoV-19/England/BRIS-122483/2020\|EPI_ISL_440777\|20 | ···················· | ···················· | ····················· | ···················· | ···················· | ···················· |
| B | 7662 | hCoV-19/England/BRIS-122C7C/2020\|EPI_ISL_440779\|20 | ···················· | ···················· | ····················· | ···················· | ···················· | ···················· |
| B | 7663 | hCoV-19/England/BRIS-1229DF/2020\|EPI_ISL_440781\|20 | ···················· | ···················· | ····················· | ···················· | ···················· | ···················· |
| B | 7664 | hCoV-19/England/BRIS-122EA3/2020\|EPI_ISL_440783\|20 | ···················· | ···················· | ····················· | ···················· | ···················· | ···················· |
| B | 7665 | hCoV-19/England/BRIS-122A81/2020\|EPI_ISL_440784\|20 | ···················· | ···················· | ····················· | ···················· | ···················· | ···················· |
| B | 7666 | hCoV-19/England/BRIS-122BBB/2020\|EPI_ISL_440785\|20 | ···················· | ···················· | ····················· | ···················· | ···················· | ···················· |
| B | 7667 | hCoV-19/England/BRIS-122E2B/2020\|EPI_ISL_440787\|20 | ···················· | ···················· | ····················· | ···················· | ···················· | ···················· |
| B | 7668 | hCoV-19/England/BRIS-122605/2020\|EPI_ISL_440788\|20 | ···················· | ···················· | ····················· | ···················· | ···················· | ···················· |
| B | 7669 | hCoV-19/England/BRIS-122D1F/2020\|EPI_ISL_440789\|20 | ···················· | ···················· | ····················· | ···················· | ···················· | ···················· |
| B | 7670 | hCoV-19/England/BRIS-1225F9/2020\|EPI_ISL_440790\|20 | ···················· | ···················· | ····················· | ···················· | ···················· | ···················· |
| B | 7671 | hCoV-19/England/BRIS-122799/2020\|EPI_ISL_440791\|20 | ···················· | ···················· | ····················· | ···················· | ···················· | ···················· |
| B | 7672 | hCoV-19/England/BRIS-122C4F/2020\|EPI_ISL_440793\|20 | ···················· | ···················· | ····················· | ···················· | ···················· | ···················· |
| B | 7673 | hCoV-19/England/BRIS-1224CF/2020\|EPI_ISL_440794\|20 | ···················· | ···················· | ····················· | ···················· | ···················· | ···················· |
| B | 7674 | hCoV-19/England/BRIS-1223B3/2020\|EPI_ISL_440795\|20 | ···················· | ···················· | ····················· | ···················· | ···················· | ···················· |
| B | 7675 | hCoV-19/England/BRIS-1226BA/2020\|EPI_ISL_440796\|20 | ···················· | ···················· | ····················· | ···················· | ···················· | ···················· |
| B | 7676 | hCoV-19/England/BRIS-122DF1/2020\|EPI_ISL_440799\|20 | ···················· | ···················· | ····················· | ···················· | ···················· | ···················· |
| B | 7677 | hCoV-19/Wales/PHWC-2A923/2020\|EPI_ISL_445637\|2020- | ···················· | ···················· | ····················· | ···················· | ···················· | ···················· |
| B | 7678 | hCoV-19/Wales/PHWC-2A932/2020\|EPI_ISL_445638\|2020- | ···················· | ···················· | ····················· | ···················· | ···················· | ···················· |
| B | 7679 | hCoV-19/Wales/PHWC-2A941/2020\|EPI_ISL_445639\|2020- | ···················· | ···················· | ····················· | ···················· | ···················· | ···················· |
| B | 7680 | hCoV-19/Wales/PHWC-3474A/2020\|EPI_ISL_446968\|2020- | ···················· | ···················· | ····················· | ···················· | ···················· | ···················· |
| B | 7681 | hCoV-19/India/TG-GMC-RK100/2020\|EPI_ISL_431101\|202 | ···················· | ···················· | ····················· | ···················· | ···················· | ···················· |
| B | 7682 | hCoV-19/India/TG-GMC-KN318/2020\|EPI_ISL_431102\|202 | ···················· | ···················· | ····················· | ···················· | ···················· | ···················· |
| B | 7683 | hCoV-19/Wales/PHWC-27D2A/2020\|EPI_ISL_432236\|2020- | ···················· | ···················· | ····················· | ···················· | ···················· | ···················· |
| B | 7684 | hCoV-19/Wales/PHWC-2A03D/2020\|EPI_ISL_432237\|2020- | ···················· | ···················· | ····················· | ···················· | ···················· | ···················· |
| B | 7685 | hCoV-19/Wales/PHWC-27C1E/2020\|EPI_ISL_432238\|2020- | ···················· | ···················· | ····················· | ···················· | ···················· | ···················· |
| B | 7686 | hCoV-19/Wales/PHWC-27D39/2020\|EPI_ISL_432239\|2020- | ···················· | ···················· | ····················· | ···················· | ···················· | ···················· |
| B | 7687 | hCoV-19/Wales/PHWC-27BA8/2020\|EPI_ISL_432240\|2020- | ···················· | ···················· | ····················· | ···················· | ···················· | ···················· |
| B | 7688 | hCoV-19/Wales/PHWC-2B306/2020\|EPI_ISL_432241\|2020- | ···················· | ···················· | ····················· | ···················· | ···················· | ···················· |
| B | 7689 | hCoV-19/Wales/PHWC-27328/2020\|EPI_ISL_432242\|2020- | ···················· | ····A··············· | ····················· | ···················· | ···················· | ···················· |
| B | 7690 | hCoV-19/Wales/PHWC-29BF1/2020\|EPI_ISL_432243\|2020- | ···················· | ···················· | ····················· | ···················· | ···················· | ···················· |
| B | 7691 | hCoV-19/Wales/PHWC-29D55/2020\|EPI_ISL_432244\|2020- | ···················· | ···················· | ····················· | ···················· | ···················· | ···················· |
| B | 7692 | hCoV-19/Wales/PHWC-27A23/2020\|EPI_ISL_432245\|2020- | ···················· | ···················· | ····················· | ···················· | ···················· | ···················· |
| B | 7693 | hCoV-19/Wales/PHWC-2B1FD/2020\|EPI_ISL_432246\|2020- | ···················· | ···················· | ····················· | ···················· | ···················· | ···················· |
| B | 7694 | hCoV-19/Wales/PHWC-29C85/2020\|EPI_ISL_432247\|2020- | ···················· | ···················· | ····················· | ···················· | ···················· | ···················· |
| B | 7695 | hCoV-19/Wales/PHWC-29D73/2020\|EPI_ISL_432248\|2020- | ···················· | ····A··············· | ····················· | ···················· | ···················· | ···················· |
| B | 7696 | hCoV-19/Wales/PHWC-29DBF/2020\|EPI_ISL_432249\|2020- | ···················· | ···················· | ····················· | ···················· | ···················· | ···················· |
| B | 7697 | hCoV-19/Wales/PHWC-2722B/2020\|EPI_ISL_432250\|2020- | ···················· | ···················· | ····················· | ···················· | ···················· | ···················· |
| B | 7698 | hCoV-19/Wales/PHWC-2A088/2020\|EPI_ISL_432251\|2020- | ···················· | ···················· | ····················· | ···················· | ···················· | ···················· |
| B | 7699 | hCoV-19/Wales/PHWC-2A0F1/2020\|EPI_ISL_432252\|2020- | ···················· | ···················· | ····················· | ···················· | ···················· | ···················· |
| B | 7700 | hCoV-19/Wales/PHWC-29B79/2020\|EPI_ISL_432253\|2020- | ···················· | ···················· | ····················· | ···················· | ···················· | ···················· |
| B | 7701 | hCoV-19/Wales/PHWC-2748F/2020\|EPI_ISL_432254\|2020- | ···················· | ···················· | ····················· | ···················· | ···················· | ···················· |
| B | 7702 | hCoV-19/Wales/PHWC-27531/2020\|EPI_ISL_432255\|2020- | ···················· | ···················· | ····················· | ···················· | ···················· | ···················· |
| B | 7703 | hCoV-19/Wales/PHWC-275F5/2020\|EPI_ISL_432256\|2020- | ···················· | ···················· | ····················· | ···················· | ···················· | ···················· |
| B | 7704 | hCoV-19/Wales/PHWC-2762F/2020\|EPI_ISL_432257\|2020- | ···················· | ···················· | ····················· | ···················· | ···················· | ···················· |
| B | 7705 | hCoV-19/Wales/PHWC-2757D/2020\|EPI_ISL_432258\|2020- | ···················· | ···················· | ····················· | ···················· | ···················· | ···················· |
| B | 7706 | hCoV-19/Wales/PHWC-29DFB/2020\|EPI_ISL_432259\|2020- | ···················· | ····A··············· | ····················· | ···················· | ···················· | ···················· |
| B | 7707 | hCoV-19/Wales/PHWC-27197/2020\|EPI_ISL_432260\|2020- | ···················· | ···················· | ····················· | ···················· | ···················· | ···················· |
| B | 7708 | hCoV-19/Wales/PHWC-2B1A2/2020\|EPI_ISL_432261\|2020- | ···················· | ···················· | ····················· | ···················· | ···················· | ···················· |
| B | 7709 | hCoV-19/Wales/PHWC-2720D/2020\|EPI_ISL_432262\|2020- | ···················· | ···················· | ····················· | ···················· | ···················· | ···················· |
| B | 7710 | hCoV-19/Wales/PHWC-2B00F/2020\|EPI_ISL_432263\|2020- | ···················· | ···················· | ····················· | ···················· | ···················· | ···················· |
| B | 7711 | hCoV-19/Wales/PHWC-2B148/2020\|EPI_ISL_432264\|2020- | ···················· | ···················· | ····················· | ···················· | ···················· | ···················· |
| B | 7712 | hCoV-19/Wales/PHWC-2B01E/2020\|EPI_ISL_432265\|2020- | ···················· | ···················· | ····················· | ···················· | ···················· | ···················· |
| B | 7713 | hCoV-19/Wales/PHWC-27513/2020\|EPI_ISL_432266\|2020- | ···················· | ···················· | ····················· | ···················· | ···················· | ···················· |
| B | 7714 | hCoV-19/Wales/PHWC-29E07/2020\|EPI_ISL_432267\|2020- | ···················· | ···················· | ····················· | ···················· | ···················· | ···················· |
| B | 7715 | hCoV-19/Wales/PHWC-2B0D2/2020\|EPI_ISL_432268\|2020- | ···················· | ···················· | ····················· | ···················· | ···················· | ···················· |
| B | 7716 | hCoV-19/Wales/PHWC-2AE42/2020\|EPI_ISL_432269\|2020- | ···················· | ·············A······ | ····················· | ···················· | ···················· | ···················· |
| B | 7717 | hCoV-19/Wales/PHWC-27935/2020\|EPI_ISL_432270\|2020- | ···················· | ···················· | ····················· | ···················· | ···················· | ···················· |
| B | 7718 | hCoV-19/Wales/PHWC-2B04B/2020\|EPI_ISL_432271\|2020- | ···················· | ···················· | ····················· | ···················· | ···················· | ···················· |
| B | 7719 | hCoV-19/Wales/PHWC-2B1EE/2020\|EPI_ISL_432272\|2020- | ···················· | ···················· | ····················· | ···················· | ···················· | ···················· |
| B | 7720 | hCoV-19/Wales/PHWC-2B218/2020\|EPI_ISL_432273\|2020- | ···················· | ···················· | ····················· | ···················· | ···················· | ···················· |
| B | 7721 | hCoV-19/Wales/PHWC-2A11C/2020\|EPI_ISL_432274\|2020- | ···················· | ···················· | ····················· | ···················· | ···················· | ···················· |
| B | 7722 | hCoV-19/Wales/PHWC-2AE9D/2020\|EPI_ISL_432275\|2020- | ···················· | ···················· | ····················· | ···················· | ···················· | ···················· |
| B | 7723 | hCoV-19/Wales/PHWC-2AEBB/2020\|EPI_ISL_432276\|2020- | ···················· | ···················· | ····················· | ···················· | ···················· | ···················· |
| B | 7724 | hCoV-19/Wales/PHWC-27285/2020\|EPI_ISL_432277\|2020- | ···················· | ···················· | ····················· | ···················· | ···················· | ···················· |
| B | 7725 | hCoV-19/Wales/PHWC-275C8/2020\|EPI_ISL_432278\|2020- | ···················· | ···················· | ····················· | ···················· | ···················· | ···················· |
| B | 7726 | hCoV-19/Wales/PHWC-29C94/2020\|EPI_ISL_432279\|2020- | ···················· | ···················· | ····················· | ···················· | ···················· | ···················· |
| B | 7727 | hCoV-19/Wales/PHWC-2B184/2020\|EPI_ISL_432280\|2020- | ···················· | ···················· | ····················· | ···················· | ···················· | ···················· |
| B | 7728 | hCoV-19/Wales/PHWC-2B227/2020\|EPI_ISL_432281\|2020- | ···················· | ···················· | ····················· | ···················· | ···················· | ···················· |

| | | | | | | | | |
|---|---|---|---|---|---|---|---|---|
| B | 7729 hCoV-19/Wales/PHWC-274AD/2020\|EPI_ISL_432282\|2020- | ···················· | ···················· | ···················· | ···················· | ···················· | ···················· | |
| B | 7730 hCoV-19/Wales/PHWC-2B05A/2020\|EPI_ISL_432283\|2020- | ···················· | ·A·················· | ···················· | ···················· | ···················· | ···················· | |
| B | 7731 hCoV-19/Wales/PHWC-2B0B4/2020\|EPI_ISL_432284\|2020- | ···················· | ···················· | ···················· | ···················· | ···················· | ···················· | |
| B | 7732 hCoV-19/Wales/PHWC-271E2/2020\|EPI_ISL_432285\|2020- | ···················· | ···A················ | ···················· | ···················· | ···················· | ···················· | |
| B | 7733 hCoV-19/Wales/PHWC-2B236/2020\|EPI_ISL_432286\|2020- | ···················· | ···A················ | ···················· | ···················· | ···················· | ···················· | |
| B | 7734 hCoV-19/Wales/PHWC-2772C/2020\|EPI_ISL_432287\|2020- | ···················· | ···················· | ···················· | ···················· | ···················· | ···················· | |
| B | 7735 hCoV-19/Wales/PHWC-277A4/2020\|EPI_ISL_432288\|2020- | ···················· | ···················· | ···················· | ···················· | ···················· | ···················· | |
| B | 7736 hCoV-19/Wales/PHWC-27971/2020\|EPI_ISL_432289\|2020- | ···················· | ···················· | ···················· | ···················· | ···················· | ···················· | |
| B | 7737 hCoV-19/Wales/PHWC-2730A/2020\|EPI_ISL_432290\|2020- | ···················· | ···················· | ···················· | ···················· | ···················· | ···················· | |
| B | 7738 hCoV-19/Wales/PHWC-2B2AF/2020\|EPI_ISL_432291\|2020- | ···················· | ···A················ | ···················· | ···················· | ···················· | ···················· | |
| B | 7739 hCoV-19/Wales/PHWC-29C58/2020\|EPI_ISL_432292\|2020- | ···················· | ···················· | ···················· | ···················· | ···················· | ···················· | |
| B | 7740 hCoV-19/Wales/PHWC-27504/2020\|EPI_ISL_432293\|2020- | ···················· | ···················· | ···················· | ···················· | ···················· | ···················· | |
| B | 7741 hCoV-19/Wales/PHWC-29FD7/2020\|EPI_ISL_432294\|2020- | ···················· | ···················· | ···················· | ···················· | ···················· | ···················· | |
| B | 7742 hCoV-19/Wales/PHWC-2AF9A/2020\|EPI_ISL_432295\|2020- | ···················· | ···················· | ···················· | ···················· | ···················· | ···················· | |
| B | 7743 hCoV-19/Wales/PHWC-2B078/2020\|EPI_ISL_432296\|2020- | ···················· | ···················· | ···················· | ···················· | ···················· | ···················· | |
| B | 7744 hCoV-19/Wales/PHWC-29D82/2020\|EPI_ISL_432297\|2020- | ···················· | ···················· | ···················· | ···················· | ···················· | ···················· | |
| B | 7745 hCoV-19/Wales/PHWC-27C0F/2020\|EPI_ISL_432298\|2020- | ···················· | ···················· | ···················· | ···················· | ···················· | ···················· | |
| B | 7746 hCoV-19/Wales/PHWC-27470/2020\|EPI_ISL_432299\|2020- | ···················· | ···················· | ···················· | ···················· | ···················· | ···················· | |
| B | 7747 hCoV-19/Wales/PHWC-27C4B/2020\|EPI_ISL_432416\|2020- | ···················· | ···················· | ···················· | ···················· | ···················· | ···················· | |
| B | 7748 hCoV-19/Wales/PHWC-2872B/2020\|EPI_ISL_432417\|2020- | ···················· | ···················· | ···················· | ···················· | ···················· | ···················· | |
| B | 7749 hCoV-19/Wales/PHWC-2B1B1/2020\|EPI_ISL_432418\|2020- | ···················· | ···················· | ···················· | ···················· | ···················· | ···················· | |
| B | 7750 hCoV-19/Wales/PHWC-27EEB/2020\|EPI_ISL_432419\|2020- | ···················· | ···A················ | ···················· | ···················· | ···················· | ···················· | |
| B | 7751 hCoV-19/Wales/PHWC-285B8/2020\|EPI_ISL_432421\|2020- | ···················· | ···················· | ···················· | ···················· | ···················· | ···················· | |
| B | 7752 hCoV-19/Wales/PHWC-273EC/2020\|EPI_ISL_432423\|2020- | ···················· | ···················· | ···················· | ···················· | ···················· | ···················· | |
| B | 7753 hCoV-19/Wales/PHWC-28600/2020\|EPI_ISL_432425\|2020- | ···················· | ···················· | ···················· | ···················· | ···················· | ···················· | |
| B | 7754 hCoV-19/Wales/PHWC-281C3/2020\|EPI_ISL_432428\|2020- | ···················· | ···················· | ···················· | ···················· | ···················· | ···················· | |
| B | 7755 hCoV-19/Wales/PHWC-28749/2020\|EPI_ISL_432430\|2020- | ···················· | ···················· | ···················· | ···················· | ···················· | ···················· | |
| B | 7756 hCoV-19/Scotland/CVR1398/2020\|EPI_ISL_433567\|2020- | ···················· | ·A·················· | ···················· | ···················· | ···················· | ···················· | |
| B | 7757 hCoV-19/Scotland/CVR1400/2020\|EPI_ISL_433568\|2020- | ···················· | ···················· | ···················· | ···················· | ···················· | ···················· | |
| B | 7758 hCoV-19/Scotland/CVR1403/2020\|EPI_ISL_433569\|2020- | ···················· | ···················A | ···················· | ···················· | ···················· | ···················· | 1 |
| B | 7759 hCoV-19/Scotland/CVR1404/2020\|EPI_ISL_433570\|2020- | ···················· | ·A·················· | ···················· | ···················· | ···················· | ···················· | |
| B | 7760 hCoV-19/Scotland/CVR1406/2020\|EPI_ISL_433571\|2020- | ···················· | ···················· | ···················· | ···················· | ···················· | ···················· | |
| B | 7761 hCoV-19/Scotland/CVR1407/2020\|EPI_ISL_433572\|2020- | ···················· | ···················· | ···················· | ···················· | ···················· | ···················· | |
| B | 7762 hCoV-19/Scotland/CVR1410/2020\|EPI_ISL_433574\|2020- | ···················· | ···················· | ···················· | ···················· | ···················· | ···················· | |
| B | 7763 hCoV-19/Scotland/CVR1412/2020\|EPI_ISL_433575\|2020- | ···················· | ···················· | ···················· | ···················· | ···················· | ···················· | |
| B | 7764 hCoV-19/Scotland/CVR1414/2020\|EPI_ISL_433576\|2020- | ···················· | ·A·················· | ···················· | ···················· | ···················· | ···················· | |
| B | 7765 hCoV-19/Scotland/CVR1426/2020\|EPI_ISL_433580\|2020- | ···················· | ···················· | ···················· | ···················· | ···················· | ···················· | |
| B | 7766 hCoV-19/Scotland/CVR1429/2020\|EPI_ISL_433581\|2020- | ···················· | ·A·················· | ···················· | ···················· | ···················· | ···················· | |
| B | 7767 hCoV-19/Scotland/CVR1430/2020\|EPI_ISL_433582\|2020- | ···················· | ···················· | ···················· | ···················· | ···················· | ···················· | |
| B | 7768 hCoV-19/Scotland/CVR1431/2020\|EPI_ISL_433583\|2020- | ···················· | ···················· | ···················· | ···················· | ···················· | ···················· | |
| B | 7769 hCoV-19/Scotland/CVR1432/2020\|EPI_ISL_433584\|2020- | ···················· | ···················· | ···················· | ···················· | ···················· | ···················· | |
| B | 7770 hCoV-19/Scotland/CVR1434/2020\|EPI_ISL_433585\|2020- | ···················· | ···················· | ···················· | ···················· | ···················· | ···················· | |
| B | 7771 hCoV-19/Scotland/CVR1437/2020\|EPI_ISL_433586\|2020- | ···················· | ···················· | ···················· | ···················· | ···················· | ···················· | |
| B | 7772 hCoV-19/Scotland/CVR1438/2020\|EPI_ISL_433587\|2020- | ···················· | ···················· | ···················· | ···················· | ···················· | ···················· | |
| B | 7773 hCoV-19/Scotland/CVR1443/2020\|EPI_ISL_433588\|2020- | ···················· | ··············C····· | ···················· | ···················· | ···················· | ···················· | |
| B | 7774 hCoV-19/Scotland/CVR1444/2020\|EPI_ISL_433589\|2020- | ···················· | ·A·················· | ···················· | ···················· | ···················· | ···················· | |
| B | 7775 hCoV-19/Scotland/CVR1450/2020\|EPI_ISL_433590\|2020- | ···················· | ·A·················· | ···················· | ···················· | ···················· | ···················· | |
| B | 7776 hCoV-19/Scotland/CVR1452/2020\|EPI_ISL_433591\|2020- | ···················· | ···················· | ···················· | ···················· | ···················· | ···················· | |
| B | 7777 hCoV-19/Scotland/CVR1468/2020\|EPI_ISL_433593\|2020- | ···················· | ···················A | ···················· | ···················· | ···················· | ···················· | 1 |
| B | 7778 hCoV-19/Scotland/CVR1469/2020\|EPI_ISL_433594\|2020- | ···················· | ···················· | ···················· | ···················· | ···················· | ···················· | |
| B | 7779 hCoV-19/Scotland/CVR1470/2020\|EPI_ISL_433595\|2020- | ···················· | ···················· | ···················· | ···················· | ···················· | ···················· | |
| B | 7780 hCoV-19/Scotland/CVR1472/2020\|EPI_ISL_433597\|2020- | ···················· | ···················· | ···················· | ···················· | ···················· | ···················· | |
| B | 7781 hCoV-19/Scotland/CVR1474/2020\|EPI_ISL_433598\|2020- | ···················· | ···················A | ···················· | ···················· | ···················· | ···················· | 1 |
| B | 7782 hCoV-19/England/CAMB-784E9/2020\|EPI_ISL_433743\|202 | ···················· | ···················· | ···················· | ···················· | ···················· | ···················· | |
| B | 7783 hCoV-19/England/CAMB-78504/2020\|EPI_ISL_433744\|202 | ···················· | ···················· | ···················· | ···················· | ···················· | ···················· | |
| B | 7784 hCoV-19/England/CAMB-78531/2020\|EPI_ISL_433745\|202 | ···················· | ···················· | ···················· | ···················· | ···················· | ···················· | |
| B | 7785 hCoV-19/England/CAMB-78540/2020\|EPI_ISL_433746\|202 | ···················· | ···················· | ···················· | ···················· | ···················· | ···················· | |
| B | 7786 hCoV-19/England/CAMB-7855F/2020\|EPI_ISL_433747\|202 | ···················· | ···················· | ···················· | ···················· | ···················· | ···················· | |
| B | 7787 hCoV-19/England/CAMB-7856E/2020\|EPI_ISL_433748\|202 | ···················· | ···················· | ···················· | ···················· | ···················· | ···················· | |
| B | 7789 hCoV-19/England/CAMB-7858C/2020\|EPI_ISL_433750\|202 | ···················· | ···················· | ···················· | ···················· | ···················· | ···················· | |
| B | 7790 hCoV-19/England/CAMB-7859B/2020\|EPI_ISL_433751\|202 | ···················· | ···················· | ···················· | ···················· | ···················· | ···················· | |
| B | 7791 hCoV-19/England/CAMB-785AA/2020\|EPI_ISL_433752\|202 | ···················· | ···················· | ···················· | ···················· | ···················· | ···················· | |
| B | 7792 hCoV-19/England/CAMB-785C8/2020\|EPI_ISL_433753\|202 | ···················· | ···················· | ···················· | ···················· | ···················· | ···················· | |
| B | 7793 hCoV-19/England/CAMB-785F5/2020\|EPI_ISL_433755\|202 | ···················· | ···················· | ···················· | ···················· | ···················· | ···················· | |
| B | 7794 hCoV-19/England/CAMB-7862F/2020\|EPI_ISL_433757\|202 | ···················· | ···················· | ···················· | ···················· | ···················· | ···················· | |
| B | 7795 hCoV-19/England/CAMB-7864D/2020\|EPI_ISL_433758\|202 | ···················· | ···················· | ···················· | ···················· | ···················· | ···················· | |
| B | 7796 hCoV-19/England/CAMB-7866B/2020\|EPI_ISL_433759\|202 | ···················· | ···················A | ···················· | ···················· | ···················· | ···················· | 1 |
| B | 7797 hCoV-19/England/CAMB-7867A/2020\|EPI_ISL_433760\|202 | ···················· | ···················· | ···················· | ···················· | ···················· | ···················· | |
| B | 7798 hCoV-19/England/CAMB-78689/2020\|EPI_ISL_433761\|202 | ···················· | ···················· | ···················· | ···················· | ···················· | ···················· | |
| B | 7799 hCoV-19/England/CAMB-786A7/2020\|EPI_ISL_433762\|202 | ···················· | ···················· | ···················· | ···················· | ···················· | ···················· | |
| B | 7800 hCoV-19/England/CAMB-786C5/2020\|EPI_ISL_433763\|202 | ···················· | ···················· | ···················· | ···················· | ···················· | ···················· | |
| B | 7801 hCoV-19/England/CAMB-786E3/2020\|EPI_ISL_433764\|202 | ···················· | ···················· | ···················· | ···················· | ···················· | ···················· | |
| B | 7802 hCoV-19/USA/TX-HMH0269/2020\|EPI_ISL_434898\|2020-03 | ···················· | ···················· | ···················· | ···················· | ···················· | ···················· | |
| B | 7803 hCoV-19/USA/TX-HMH0271/2020\|EPI_ISL_434899\|2020-03 | ···················· | ···················· | ···················· | ···················· | ···················· | ···················· | |
| B | 7804 hCoV-19/USA/MI-MDHHS-SC20013/2020\|EPI_ISL_437100\|2 | ···················· | ···················· | ···················· | ···················· | ···················· | ···················· | |
| B | 7805 hCoV-19/USA/MI-MDHHS-SC20014/2020\|EPI_ISL_437101\|2 | ···················· | ···················· | ···················· | ···················· | ···················· | ···················· | |
| B | 7806 hCoV-19/USA/MI-MDHHS-SC20020/2020\|EPI_ISL_437107\|2 | ···················· | ···················· | ···················· | ···················· | ···················· | ···················· | |
| B | 7807 hCoV-19/England/CAMB-75D4B/2020\|EPI_ISL_438420\|202 | ···················· | ···················· | ···················· | ···················· | ···················· | ···················· | |

| | | | | | | | |
|---|---|---|---|---|---|---|---|
| B | 7808 hCoV-19/England/CAMB-766D6/2020\|EPI_ISL_438421\|202 | ···················· | ···················· | ····················· | ···················· | ···················· | ···················· |
| B | 7809 hCoV-19/England/CAMB-76B6E/2020\|EPI_ISL_438422\|202 | ···················· | ···················· | ····················· | ···················· | ···················· | ···················· |
| B | 7810 hCoV-19/England/CAMB-75D2D/2020\|EPI_ISL_438423\|202 | ···················· | ···················· | ····················· | ···················· | ···················· | ···················· |
| B | 7811 hCoV-19/England/CAMB-76991/2020\|EPI_ISL_438424\|202 | ···················· | ···················· | ····················· | ···················· | ···················· | ···················· |
| B | 7812 hCoV-19/England/CAMB-7673D/2020\|EPI_ISL_438425\|202 | ···················· | ···················· | ····················· | ···················· | ···················· | ···················· |
| B | 7813 hCoV-19/England/CAMB-764CD/2020\|EPI_ISL_438429\|202 | ···················· | ···················· | ····················· | ···················· | ···················· | ···················· |
| B | 7814 hCoV-19/England/CAMB-75CB7/2020\|EPI_ISL_438430\|202 | ···················· | ···················· | ····················· | ···················· | ···················· | ···················· |
| B | 7815 hCoV-19/England/CAMB-76BD7/2020\|EPI_ISL_438431\|202 | ···················· | ···················· | ····················· | ···················· | ···················· | ···················· |
| B | 7816 hCoV-19/England/CAMB-76B04/2020\|EPI_ISL_438434\|202 | ···················· | ···················· | ····················· | ···················· | ···················· | ···················· |
| B | 7817 hCoV-19/England/CAMB-7664F/2020\|EPI_ISL_438436\|202 | ···················· | ···················· | ····················· | ···················· | ···················· | ···················· |
| B | 7818 hCoV-19/England/CAMB-762A5/2020\|EPI_ISL_438437\|202 | ···················· | ···················· | ····················· | ···················· | ···················· | ···················· |
| B | 7819 hCoV-19/England/LIVE-9927F/2020\|EPI_ISL_439704\|202 | ···················· | ···················· | ····················· | ···················· | ···················· | ···················· |
| B | 7820 hCoV-19/England/LIVE-99707/2020\|EPI_ISL_439751\|202 | ···················· | ···················· | ····················· | ···················· | ···················· | ···················· |
| B | 7821 hCoV-19/England/LIVE-99716/2020\|EPI_ISL_439752\|202 | ···················· | ···················· | ····················· | ···················· | ···················· | ···················· |
| B | 7822 hCoV-19/England/LIVE-99725/2020\|EPI_ISL_439753\|202 | ···················· | ···················· | ····················· | ···················· | ···················· | ···················· |
| B | 7823 hCoV-19/England/LIVE-99743/2020\|EPI_ISL_439754\|202 | ···················· | ···················· | ····················· | ···················· | ···················· | ···················· |
| B | 7824 hCoV-19/England/LIVE-99761/2020\|EPI_ISL_439756\|202 | ···················· | ···················· | ····················· | ···················· | ···················· | ···················· |
| B | 7825 hCoV-19/England/LIVE-9978F/2020\|EPI_ISL_439757\|202 | ···················· | ···················· | ····················· | ···················· | ···················· | ···················· |
| B | 7826 hCoV-19/England/LIVE-997E9/2020\|EPI_ISL_439762\|202 | ···················· | ···················· | ····················· | ···················· | ···················· | ···················· |
| B | 7827 hCoV-19/England/LIVE-997F8/2020\|EPI_ISL_439763\|202 | ···················· | ···················· | ····················· | ···················· | ···················· | ···················· |
| B | 7828 hCoV-19/England/LIVE-99840/2020\|EPI_ISL_439765\|202 | ···················· | ···················· | ····················· | ···················· | ···················· | ···················· |
| B | 7829 hCoV-19/England/LIVE-9986E/2020\|EPI_ISL_439767\|202 | ···················· | ···················· | ····················· | ···················· | ···················· | ···················· |
| B | 7830 hCoV-19/England/BRIS-122C21/2020\|EPI_ISL_440730\|20 | ···················· | ···················· | ····················· | ···················· | ···················· | ···················· |
| B | 7831 hCoV-19/England/BRIS-122571/2020\|EPI_ISL_440732\|20 | ···················· | ···················· | ····················· | ···················· | ···················· | ···················· |
| B | 7832 hCoV-19/England/BRIS-122CE5/2020\|EPI_ISL_440733\|20 | ···················· | ···················· | ····················· | ···················· | ···················· | ···················· |
| B | 7833 hCoV-19/England/BRIS-122B8E/2020\|EPI_ISL_440734\|20 | ···················· | ···················· | ····················· | ···················· | ···················· | ···················· |
| B | 7834 hCoV-19/England/BRIS-122878/2020\|EPI_ISL_440735\|20 | ···················· | ···················· | ····················· | ···················· | ···················· | ···················· |
| B | 7835 hCoV-19/England/BRIS-122BAC/2020\|EPI_ISL_440738\|20 | ···················· | ···················· | ····················· | ···················· | ···················· | ···················· |
| B | 7836 hCoV-19/England/BRIS-12285A/2020\|EPI_ISL_440740\|20 | ···················· | ···················· | ····················· | ···················· | ···················· | ···················· |
| B | 7837 hCoV-19/England/BRIS-1229EE/2020\|EPI_ISL_440741\|20 | ···················· | ···················· | ····················· | ···················· | ···················· | ···················· |
| B | 7838 hCoV-19/England/BRIS-122B7F/2020\|EPI_ISL_440742\|20 | ···················· | ···················· | ····················· | ···················· | ···················· | ···················· |
| B | 7839 hCoV-19/England/BRIS-123006/2020\|EPI_ISL_440744\|20 | ···················· | ···················· | ····················· | ···················· | ···················· | ···················· |
| B | 7840 hCoV-19/England/BRIS-122395/2020\|EPI_ISL_440745\|20 | ···················· | ···················· | ····················· | ···················· | ···················· | ···················· |
| B | 7841 hCoV-19/England/BRIS-122FCE/2020\|EPI_ISL_440746\|20 | ···················· | ···················· | ····················· | ···················· | ···················· | ···················· |
| B | 7842 hCoV-19/England/BRIS-1223A4/2020\|EPI_ISL_440747\|20 | ···················· | ···················· | ····················· | ···················· | ···················· | ···················· |
| B | 7843 hCoV-19/England/LOND-D53A1/2020\|EPI_ISL_444215\|202 | ···················· | ···················· | ····················· | ···················· | ···················· | ···················· |
| B | 7844 hCoV-19/England/LOND-D53CF/2020\|EPI_ISL_444216\|202 | ···················· | ···················· | ····················· | ···················· | ···················· | ···················· |
| B | 7845 hCoV-19/England/LOND-D53DE/2020\|EPI_ISL_444217\|202 | ···················· | ···················· | ····················· | ···················· | ···················· | ···················· |
| B | 7846 hCoV-19/England/LOND-D5417/2020\|EPI_ISL_444218\|202 | ···················· | ···················· | ····················· | ···················· | ···················· | ···················· |
| B | 7847 hCoV-19/England/LOND-D5426/2020\|EPI_ISL_444219\|202 | ···T················ | ···················· | ····················· | ···················· | ···················· | ···················· |
| B | 7848 hCoV-19/England/LOND-D5435/2020\|EPI_ISL_444220\|202 | ···················· | ···················· | ····················· | ···················· | ···················· | ···················· |
| B | 7849 hCoV-19/England/LOND-D5444/2020\|EPI_ISL_444221\|202 | ···················· | ·N·················· | ····················· | ···················· | ···················· | ···················· |
| B | 7850 hCoV-19/England/LOND-D5453/2020\|EPI_ISL_444222\|202 | ···················· | ···················· | ····················· | ···················· | ···················· | ···················· |
| B | 7851 hCoV-19/England/LOND-D5471/2020\|EPI_ISL_444223\|202 | ···················· | ···················· | ····················· | ···················· | ···················· | ···················· |
| B | 7852 hCoV-19/England/LOND-D5480/2020\|EPI_ISL_444224\|202 | ···················· | ···················· | ····················· | ···················· | ···················· | ···················· |
| B | 7853 hCoV-19/England/LOND-D549F/2020\|EPI_ISL_444225\|202 | ···················· | ···················· | ····················· | ···················· | ···················· | ···················· |
| B | 7854 hCoV-19/England/LOND-D54AE/2020\|EPI_ISL_444226\|202 | ···················· | ···················· | ····················· | ···················· | ···················· | ···················· |
| B | 7855 hCoV-19/England/LOND-D54DB/2020\|EPI_ISL_444227\|202 | ···················· | ···················· | ····················· | ···················· | ···················· | ···················· |
| B | 7856 hCoV-19/England/LOND-D54F9/2020\|EPI_ISL_444228\|202 | ···················· | ···················· | ····················· | ···················· | ···················· | ···················· |
| B | 7857 hCoV-19/England/LOND-D5514/2020\|EPI_ISL_444229\|202 | ···················· | ···················· | ····················· | ···················· | ···················· | ···················· |
| B | 7858 hCoV-19/England/LOND-D5523/2020\|EPI_ISL_444230\|202 | ···················· | ···················· | ····················· | ···················· | ···················· | ···················· |
| B | 7859 hCoV-19/England/LOND-D5532/2020\|EPI_ISL_444231\|202 | ···················· | ···················· | ····················· | ···················· | ···················· | ···················· |
| B | 7860 hCoV-19/England/LOND-D5541/2020\|EPI_ISL_444232\|202 | ···················· | ···················· | ····················· | ···················· | ···················· | ···················· |
| B | 7861 hCoV-19/England/LOND-D5550/2020\|EPI_ISL_444233\|202 | ···················· | ···················· | ····················· | ···················· | ···················· | ···················· |
| B | 7862 hCoV-19/England/LOND-D557E/2020\|EPI_ISL_444235\|202 | ···················· | ···················· | ····················· | ···················· | ···················· | ···················· |
| B | 7863 hCoV-19/England/LOND-D5B5E/2020\|EPI_ISL_444236\|202 | ···················· | ···················· | ····················· | ···················· | ···················· | ···················· |
| B | 7864 hCoV-19/England/LOND-D5B9A/2020\|EPI_ISL_444238\|202 | ···················· | ···················· | ····················· | ···················· | ···················· | ···················· |
| B | 7865 hCoV-19/England/LOND-D5BC7/2020\|EPI_ISL_444239\|202 | ···················· | ···················· | ····················· | ···················· | ···················· | ···················· |
| B | 7866 hCoV-19/England/LOND-D5CB5/2020\|EPI_ISL_444241\|202 | ···················· | ···················· | ····················· | ···················· | ···················· | ···················· |
| B | 7867 hCoV-19/England/LOND-D5CD3/2020\|EPI_ISL_444242\|202 | ···················· | ···················· | ····················· | ···················· | ···················· | ···················· |
| B | 7868 hCoV-19/England/LOND-D5D3A/2020\|EPI_ISL_444243\|202 | ···················· | ···················· | ····················· | ···················· | ···················· | ···················· |
| B | 7869 hCoV-19/England/LOND-D5D49/2020\|EPI_ISL_444244\|202 | ···················· | ···················· | ····················· | ···················· | ···················· | ···················· |
| B | 7870 hCoV-19/England/LOND-D5D58/2020\|EPI_ISL_444245\|202 | ···················· | ···················· | ····················· | ···················· | ···················· | ···················· |
| B | 7871 hCoV-19/England/LOND-D5D67/2020\|EPI_ISL_444246\|202 | ···················· | ···················· | ····················· | ···················· | ···················· | ···················· |
| B | 7872 hCoV-19/England/LOND-D5DA3/2020\|EPI_ISL_444247\|202 | ···················· | ···················· | ····················· | ···················· | ···················· | ···················· |
| B | 7873 hCoV-19/England/LOND-D5DEF/2020\|EPI_ISL_444248\|202 | ···················· | ···················· | ····················· | ···················· | ···················· | ···················· |
| B | 7874 hCoV-19/England/LOND-D5E19/2020\|EPI_ISL_444249\|202 | ···················· | ···················· | ····················· | ···················· | ···················· | ···················· |
| B | 7875 hCoV-19/England/LOND-D5E37/2020\|EPI_ISL_444250\|202 | ···················· | ···················· | ····················· | ···················· | ···················· | ···················· |
| B | 7876 hCoV-19/England/LOND-D5E55/2020\|EPI_ISL_444251\|202 | ···················· | ···················· | ····················· | ···················· | ···················· | ···················· |
| B | 7877 hCoV-19/England/LOND-D5E64/2020\|EPI_ISL_444252\|202 | ···················· | ···················· | ····················· | ···················· | ···················· | ···················· |
| B | 7878 hCoV-19/England/LOND-D5E82/2020\|EPI_ISL_444253\|202 | ···················· | ···················· | ····················· | ···················· | ···················· | ···················· |
| B | 7879 hCoV-19/England/LOND-D5E91/2020\|EPI_ISL_444254\|202 | ···················· | ···················· | ····················· | ···················· | ···················· | ···················· |
| B | 7880 hCoV-19/England/LOND-D5EBF/2020\|EPI_ISL_444256\|202 | ···················· | ···················· | ····················· | ···················· | ···················· | ···················· |
| B | 7881 hCoV-19/England/LOND-D5ECE/2020\|EPI_ISL_444257\|202 | ···················· | ···················· | ····················· | ···················· | ···················· | ···················· |
| B | 7882 hCoV-19/England/LOND-D5EDD/2020\|EPI_ISL_444258\|202 | ···················· | ···················· | ····················· | ···················· | ···················· | ···················· |
| B | 7883 hCoV-19/England/LOND-D5EEC/2020\|EPI_ISL_444259\|202 | ···················· | ···················· | ····················· | ···················· | ···················· | ···················· |
| B | 7884 hCoV-19/England/LOND-D5EFB/2020\|EPI_ISL_444260\|202 | ···················· | ···················· | ····················· | ···················· | ···················· | ···················· |
| B | 7885 hCoV-19/England/LOND-D5F07/2020\|EPI_ISL_444261\|202 | ···················· | ···················· | ····················· | ···················· | ···················· | ···················· |

| | | | | | | | |
|---|---|---|---|---|---|---|---|
| B | 7886 hCoV-19/England/LOND-D5F16/2020\|EPI_ISL_444262\|202 | ···················· | ···················· | ····················· | ···················· | ···················· | ···················· |
| B | 7887 hCoV-19/England/LOND-D5F25/2020\|EPI_ISL_444263\|202 | ···················· | ···················· | ····················· | ···················· | ···················· | ···················· |
| B | 7888 hCoV-19/England/LOND-D5F43/2020\|EPI_ISL_444265\|202 | ···················· | ···················· | ····················· | ···················· | ···················· | ···················· |
| B | 7889 hCoV-19/England/LOND-D5FDA/2020\|EPI_ISL_444268\|202 | ···················· | ···················· | ····················· | ···················· | ···················· | ···················· |
| B | 7890 hCoV-19/England/LOND-D5FF8/2020\|EPI_ISL_444269\|202 | ···················· | ···················· | ····················· | ···················· | ···················· | ···················· |
| B | 7891 hCoV-19/England/LOND-D6012/2020\|EPI_ISL_444270\|202 | ···················· | ···················· | ····················· | ···················· | ···················· | ···················· |
| B | 7892 hCoV-19/England/LOND-D606D/2020\|EPI_ISL_444272\|202 | ···················· | ···················· | ····················· | ···················· | ···················· | ···················· |
| B | 7893 hCoV-19/Guangzhou/IQTC05/2020\|EPI_ISL_444273\|2020- | ···················· | ···················· | ····················· | ···················· | ···················· | ···················· |
| B | 7894 hCoV-19/Taiwan/CGMH-CGU-21/2020\|EPI_ISL_444274\|202 | ···················· | ···················· | ····················· | ···················· | ···················· | ···················· |
| B | 7895 hCoV-19/Taiwan/CGMH-CGU-23/2020\|EPI_ISL_444276\|202 | ···················· | ···················· | ····················· | ···················· | ···················· | ···················· |
| B | 7896 hCoV-19/Taiwan/CGMH-CGU-24/2020\|EPI_ISL_444277\|202 | ···················· | ···················· | ····················· | ···················· | ···················· | ···················· |
| B | 7897 hCoV-19/Taiwan/CGMH-CGU-25/2020\|EPI_ISL_444278\|202 | ···················· | ···················· | ····················· | ···················· | ···················· | ···················· |
| B | 7898 hCoV-19/England/BIRM-5E2A3/2020\|EPI_ISL_444279\|202 | ···················· | ···················· | ····················· | ···················· | ···················· | ···················· |
| B | 7899 hCoV-19/England/BIRM-5E2B2/2020\|EPI_ISL_444280\|202 | ···················· | ···················· | ····················· | ···················· | ···················· | ···················· |
| B | 7900 hCoV-19/England/BIRM-5E2C1/2020\|EPI_ISL_444281\|202 | ···················· | ···················· | ····················· | ···················· | ···················· | ···················· |
| B | 7901 hCoV-19/England/BIRM-5E2EF/2020\|EPI_ISL_444283\|202 | ···················· | ···················· | ····················· | ···················· | ···················· | ···················· |
| B | 7903 hCoV-19/England/BIRM-5E30A/2020\|EPI_ISL_444285\|202 | ···················· | ···················· | ····················· | ···················· | ···················· | ···················· |
| B | 7904 hCoV-19/England/BIRM-5E319/2020\|EPI_ISL_444286\|202 | ···················· | ···················· | ····················· | ···················· | ···················· | ···················· |
| B | 7905 hCoV-19/England/BIRM-5E346/2020\|EPI_ISL_444288\|202 | ···················· | ···················· | ····················· | ···················· | ···················· | ···················· |
| B | 7906 hCoV-19/England/BIRM-5E364/2020\|EPI_ISL_444289\|202 | ···················· | ···················· | ····················· | ···················· | ···················· | ···················· |
| B | 7908 hCoV-19/England/BIRM-5E391/2020\|EPI_ISL_444291\|202 | ···················· | ···················· | ····················· | ···················· | ···················· | ···················· |
| B | 7909 hCoV-19/England/BIRM-5E3A0/2020\|EPI_ISL_444292\|202 | ···················· | ···················· | ····················· | ···················· | ···················· | ···················· |
| B | 7910 hCoV-19/England/BIRM-5E3BF/2020\|EPI_ISL_444293\|202 | ···················· | ···················· | ····················· | ···················· | ···················· | ···················· |
| B | 7912 hCoV-19/England/CAMB-7FC9F/2020\|EPI_ISL_444400\|202 | ···················· | ···················· | ····················· | ···················· | ···················· | ···················· |
| B | 7913 hCoV-19/Wales/PHWC-2A2FB/2020\|EPI_ISL_445552\|2020- | ···················· | ···················· | ····················· | ···················· | ···················· | ···················· |
| B | 7914 hCoV-19/Wales/PHWC-2A316/2020\|EPI_ISL_445553\|2020- | ···················· | ···················· | ····················· | ···················· | ···················· | ···················· |
| B | 7915 hCoV-19/Wales/PHWC-2A3DA/2020\|EPI_ISL_445564\|2020- | ···················· | ···················· | ····················· | ···················· | ···················· | ···················· |
| B | 7916 hCoV-19/Wales/PHWC-2A422/2020\|EPI_ISL_445567\|2020- | ···················· | ···················· | ····················· | ···················· | ···················· | ···················· |
| B | 7917 hCoV-19/Wales/PHWC-2A49B/2020\|EPI_ISL_445571\|2020- | ···················· | ···················· | ····················· | ···················· | ···················· | ···················· |
| B | 7918 hCoV-19/Wales/PHWC-2A4AA/2020\|EPI_ISL_445572\|2020- | ···················· | ···················· | ····················· | ···················· | ···················· | ···················· |
| B | 7919 hCoV-19/Wales/PHWC-2A4B9/2020\|EPI_ISL_445573\|2020- | ···················· | ···················· | ····················· | ···················· | ···················· | ···················· |
| B | 7920 hCoV-19/Wales/PHWC-2A4E6/2020\|EPI_ISL_445576\|2020- | ···················· | ···················· | ····················· | ···················· | ···················· | ···················· |
| B | 7921 hCoV-19/Wales/PHWC-2A55C/2020\|EPI_ISL_445583\|2020- | ···················· | ···················· | ····················· | ···················· | ···················· | ···················· |
| B | 7922 hCoV-19/Wales/PHWC-2A57A/2020\|EPI_ISL_445585\|2020- | ···················· | ···················· | ····················· | ···················· | ···················· | ···················· |
| B | 7923 hCoV-19/Wales/PHWC-2A5B6/2020\|EPI_ISL_445589\|2020- | ···················· | ···················· | ····················· | ···················· | ···················· | ···················· |
| B | 7924 hCoV-19/Wales/PHWC-2A5D4/2020\|EPI_ISL_445591\|2020- | ···················· | ····A··············· | ····················· | ···················· | ···················· | ···················· |
| B | 7925 hCoV-19/Wales/PHWC-2A60E/2020\|EPI_ISL_445594\|2020- | ···················· | ···················· | ····················· | ···················· | ···················· | ···················· |
| B | 7926 hCoV-19/Wales/PHWC-2A61D/2020\|EPI_ISL_445595\|2020- | ···················· | ···················· | ····················· | ···················· | ···················· | ···················· |
| B | 7927 hCoV-19/Wales/PHWC-2A63B/2020\|EPI_ISL_445597\|2020- | ···················· | ···················· | ····················· | ···················· | ···················· | ···················· |
| B | 7928 hCoV-19/Wales/PHWC-2B52E/2020\|EPI_ISL_445723\|2020- | ···················· | ···················· | ····················· | ···················· | ···················· | ···················· |
| B | 7929 hCoV-19/Wales/PHWC-2B53D/2020\|EPI_ISL_445724\|2020- | ···················· | ···················· | ····················· | ···················· | ···················· | ···················· |
| B | 7930 hCoV-19/Wales/PHWC-2B54C/2020\|EPI_ISL_445725\|2020- | ···················· | ···················· | ····················· | ···················· | ···················· | ···················· |
| B | 7931 hCoV-19/Wales/PHWC-2B55B/2020\|EPI_ISL_445726\|2020- | ···················· | ···················· | ····················· | ···················· | ···················· | ···················· |
| B | 7932 hCoV-19/Wales/PHWC-2B579/2020\|EPI_ISL_445727\|2020- | ···················· | ···················· | ····················· | ···················· | ···················· | ···················· |
| B | 7933 hCoV-19/Wales/PHWC-2B667/2020\|EPI_ISL_445741\|2020- | ···················· | ···················· | ····················· | ···················· | ···················· | ···················· |
| B | 7934 hCoV-19/Wales/PHWC-3415B/2020\|EPI_ISL_446877\|2020- | ···················· | ···················· | ····················· | ···················· | ···················· | ···················· |
| B | 7935 hCoV-19/Wales/PHWC-341F1/2020\|EPI_ISL_446887\|2020- | ···················· | ···················· | ····················· | ···················· | ···················· | ···················· |
| B | 7936 hCoV-19/Wales/PHWC-34285/2020\|EPI_ISL_446894\|2020- | ···················· | ···················· | ····················· | ···················· | ···················· | ···················· |
| B | 7937 hCoV-19/India/TG-GMC-KN443/2020\|EPI_ISL_431103\|202 | ···················· | ···················· | ····················· | ···················· | ···················· | ···················· |
| B | 7938 hCoV-19/India/TG-GMC-TC469/2020\|EPI_ISL_431117\|202 | ···················· | ···················· | ····················· | ···················· | ···················· | ···················· |
| B | 7939 hCoV-19/Fujian/IM3520001T/2020\|EPI_ISL_431118\|2020 | ···················· | ···················· | ····················· | ···················· | ···················· | ···················· |
| B | 7940 hCoV-19/Wales/PHWC-27F60/2020\|EPI_ISL_432400\|2020- | ···················· | ····A··············· | ····················· | ···················· | ···················· | ···················· |
| B | 7941 hCoV-19/Wales/PHWC-275B9/2020\|EPI_ISL_432401\|2020- | ···················· | ···················· | ····················· | ···················· | ···················· | ···················· |
| B | 7942 hCoV-19/Wales/PHWC-27A7E/2020\|EPI_ISL_432403\|2020- | ···················· | ···················· | ····················· | ···················· | ···················· | ···················· |
| B | 7943 hCoV-19/Wales/PHWC-24859/2020\|EPI_ISL_432405\|2020- | ···················· | ···················· | ····················· | ···················· | ···················· | ···················· |
| B | 7944 hCoV-19/Wales/PHWC-24886/2020\|EPI_ISL_432406\|2020- | ···················· | ···················· | ····················· | ···················· | ···················· | ···················· |
| B | 7945 hCoV-19/Wales/PHWC-28882/2020\|EPI_ISL_432407\|2020- | ···················· | ···················· | ····················· | ···················· | ···················· | ···················· |
| B | 7946 hCoV-19/England/SHEF-C8C2E/2020\|EPI_ISL_432458\|202 | ···················· | ···················· | ····················· | ···················· | ···················· | ···················· |
| B | 7947 hCoV-19/England/SHEF-C914C/2020\|EPI_ISL_432466\|202 | ···················· | ···················· | ····················· | ···················· | ···················· | ···················· |
| B | 7948 hCoV-19/England/CAMB-779FA/2020\|EPI_ISL_433701\|202 | ···················· | ···················· | ····················· | ···················· | ···················· | ···················· |
| B | 7949 hCoV-19/England/CAMB-78197/2020\|EPI_ISL_433702\|202 | ···················· | ···················· | ····················· | ···················· | ···················· | ···················· |
| B | 7950 hCoV-19/England/CAMB-781B5/2020\|EPI_ISL_433704\|202 | ···················· | ···················· | ····················· | ···················· | ···················· | ···················· |
| B | 7951 hCoV-19/England/CAMB-781E2/2020\|EPI_ISL_433705\|202 | ···················· | ···················· | ····················· | ···················· | ···················· | ···················· |
| B | 7952 hCoV-19/England/CAMB-7820D/2020\|EPI_ISL_433706\|202 | ···················· | ···················· | ····················· | ···················· | ···················· | ···················· |
| B | 7953 hCoV-19/England/CAMB-7822B/2020\|EPI_ISL_433707\|202 | ···················· | ···················· | ····················· | ···················· | ···················· | ···················· |
| B | 7954 hCoV-19/England/CAMB-78285/2020\|EPI_ISL_433709\|202 | ···················· | ···················· | ····················· | ···················· | ···················· | ···················· |
| B | 7955 hCoV-19/England/CAMB-78294/2020\|EPI_ISL_433710\|202 | ···················· | ···················· | ····················· | ···················· | ···················· | ···················· |
| B | 7956 hCoV-19/England/CAMB-782A3/2020\|EPI_ISL_433711\|202 | ···················· | ···················· | ····················· | ···················· | ···················· | ···················· |
| B | 7957 hCoV-19/England/CAMB-782B2/2020\|EPI_ISL_433712\|202 | ···················· | ···················· | ····················· | ···················· | ···················· | ···················· |
| B | 7958 hCoV-19/England/CAMB-782D0/2020\|EPI_ISL_433713\|202 | ···················· | ···················· | ····················· | ···················· | ···················· | ···················· |
| B | 7959 hCoV-19/England/CAMB-782EF/2020\|EPI_ISL_433714\|202 | ···················· | ···················· | ····················· | ···················· | ···················· | ···················· |
| B | 7960 hCoV-19/England/CAMB-7830A/2020\|EPI_ISL_433716\|202 | ···················· | ···················· | ····················· | ···················· | ···················· | ···················· |
| B | 7961 hCoV-19/England/CAMB-78319/2020\|EPI_ISL_433717\|202 | ···················· | ···················· | ····················· | ···················· | ···················· | ···················· |
| B | 7964 hCoV-19/England/CAMB-78346/2020\|EPI_ISL_433720\|202 | ···················· | ···················· | ····················· | ···················· | ···················· | ···················· |
| B | 7965 hCoV-19/England/CAMB-78355/2020\|EPI_ISL_433721\|202 | ···················· | ···················· | ····················· | ···················· | ···················· | ···················· |
| B | 7966 hCoV-19/England/CAMB-78364/2020\|EPI_ISL_433722\|202 | ···················· | ···················· | ····················· | ···················· | ···················· | ···················· |
| B | 7967 hCoV-19/England/CAMB-78382/2020\|EPI_ISL_433724\|202 | ···················· | ···················· | ····················· | ···················· | ···················· | ···················· |
| B | 7968 hCoV-19/England/CAMB-783DD/2020\|EPI_ISL_433728\|202 | ···················· | ···················· | ····················· | ···················· | ···················· | ···················· |

| | | | | | | | | | |
|---|---|---|---|---|---|---|---|---|---|
| B | 7969 hCoV-19/England/CAMB-783EC/2020\|EPI_ISL_433729\|202 | ···················· | ···················· | ···················· | ···················· | ···················· | ···················· | | |
| B | 7970 hCoV-19/England/CAMB-783FB/2020\|EPI_ISL_433730\|202 | ···················· | ···················· | ···················· | ···················· | ···················· | ···················· | | |
| B | 7971 hCoV-19/England/CAMB-78407/2020\|EPI_ISL_433731\|202 | ···················· | ···················· | ···················· | ···················· | ···················· | ···················· | | |
| B | 7972 hCoV-19/England/CAMB-78416/2020\|EPI_ISL_433732\|202 | ···················· | ···················· | ···················· | ···················· | ···················· | ···················· | | |
| B | 7973 hCoV-19/England/CAMB-78425/2020\|EPI_ISL_433733\|202 | ···················· | ···················· | ···················· | ···················· | ···················· | ···················· | | |
| B | 7974 hCoV-19/England/CAMB-78452/2020\|EPI_ISL_433734\|202 | ···················· | ···················· | ···················· | ···················· | ···················· | ···················· | | |
| B | 7975 hCoV-19/England/CAMB-78461/2020\|EPI_ISL_433735\|202 | ···················· | ···················· | ···················· | ···················· | ···················· | ···················· | | |
| B | 7976 hCoV-19/England/CAMB-78470/2020\|EPI_ISL_433736\|202 | ···················· | ···················· | ···················· | ···················· | ···················· | ···················· | | |
| B | 7977 hCoV-19/England/CAMB-7848F/2020\|EPI_ISL_433737\|202 | ···················· | ···················· | ···················· | ···················· | ···················· | ···················· | | |
| B | 7978 hCoV-19/England/CAMB-7849E/2020\|EPI_ISL_433738\|202 | ···················· | ···················· | ···················· | ···················· | ···················· | ···················· | | |
| B | 7979 hCoV-19/England/CAMB-784AD/2020\|EPI_ISL_433739\|202 | ···················· | ···················· | ···················· | ···················· | ···················· | ···················· | | |
| B | 7980 hCoV-19/England/CAMB-784BC/2020\|EPI_ISL_433740\|202 | ···················· | ···················· | ···················· | ···················· | ···················· | ···················· | | |
| B | 7981 hCoV-19/England/CAMB-784CB/2020\|EPI_ISL_433741\|202 | ···················· | ···················· | ···················· | ···················· | ···················· | ···················· | | |
| B | 7982 hCoV-19/England/CAMB-784DA/2020\|EPI_ISL_433742\|202 | ···················· | ···················· | ···················· | ···················· | ···················· | ···················· | | |
| B | 7983 hCoV-19/England/CAMB-7870E/2020\|EPI_ISL_433765\|202 | ···················· | ···················· | ···················· | ···················· | ···················· | ···················· | | |
| B | 7984 hCoV-19/England/CAMB-7872C/2020\|EPI_ISL_433766\|202 | ···················· | ···················· | ···················· | ···················· | ···················· | ···················· | | |
| B | 7985 hCoV-19/England/CAMB-7873B/2020\|EPI_ISL_433767\|202 | ···················· | ···················· | ···················· | ···················· | ···················· | ···················· | | |
| B | 7986 hCoV-19/England/CAMB-7874A/2020\|EPI_ISL_433768\|202 | ···················· | ···················· | ···················· | ···················· | ···················· | ···················· | | |
| B | 7987 hCoV-19/England/CAMB-78786/2020\|EPI_ISL_433770\|202 | ···················· | ···················· | ···················· | ···················· | ···················· | ···················· | | |
| B | 7988 hCoV-19/England/CAMB-787A4/2020\|EPI_ISL_433771\|202 | ···················· | ···················· | ···················· | ···················· | ···················· | ···················· | | |
| B | 7989 hCoV-19/England/CAMB-787B3/2020\|EPI_ISL_433772\|202 | ···················· | ···················· | ···················· | ···················· | ···················· | ···················· | | |
| B | 7990 hCoV-19/England/CAMB-787E0/2020\|EPI_ISL_433775\|202 | ···················· | ···················· | ···················· | ···················· | ···················· | ···················· | | |
| B | 7991 hCoV-19/England/CAMB-7880B/2020\|EPI_ISL_433776\|202 | ···················· | ···················· | ···················· | ···················· | ···················· | ···················· | | |
| B | 7992 hCoV-19/England/CAMB-7881A/2020\|EPI_ISL_433777\|202 | ···················· | ···················· | ···················· | ···················· | ···················· | ···················· | | |
| B | 7993 hCoV-19/England/CAMB-78829/2020\|EPI_ISL_433778\|202 | ···················· | ···················· | ···················· | ···················· | ···················· | ···················· | | |
| B | 7994 hCoV-19/England/CAMB-78838/2020\|EPI_ISL_433779\|202 | ···················· | ···················· | ···················· | ···················· | ···················· | ···················· | | |
| B | 7995 hCoV-19/England/CAMB-78847/2020\|EPI_ISL_433780\|202 | ···················· | ···················· | ···················· | ···················· | ···················· | ···················· | | |
| B | 7996 hCoV-19/England/CAMB-78865/2020\|EPI_ISL_433781\|202 | ···················· | ···················· | ···················· | ···················· | ···················· | ···················· | | |
| B | 7997 hCoV-19/England/CAMB-78874/2020\|EPI_ISL_433782\|202 | ···················· | ···················· | ···················· | ···················· | ···················· | ···················· | | |
| B | 7998 hCoV-19/England/CAMB-78883/2020\|EPI_ISL_433783\|202 | ···················· | ···················· | ···················· | ···················· | ···················· | ···················· | | |
| B | 7999 hCoV-19/England/CAMB-78892/2020\|EPI_ISL_433784\|202 | ···················· | ···················· | ···················· | ···················· | ···················· | ···················· | | |
| B | 8000 hCoV-19/England/CAMB-788A1/2020\|EPI_ISL_433785\|202 | ···················· | ···················· | ···················· | ···················· | ···················· | ···················· | | |
| B | 8001 hCoV-19/England/CAMB-788B0/2020\|EPI_ISL_433786\|202 | ···················· | ···················· | ···················· | ···················· | ···················· | ···················· | | |
| B | 8002 hCoV-19/England/CAMB-788CF/2020\|EPI_ISL_433787\|202 | ···················· | ···················· | ···················· | ···················· | ···················· | ···················· | | |
| B | 8003 hCoV-19/England/CAMB-788ED/2020\|EPI_ISL_433789\|202 | ···················· | ···················· | ···················· | ···················· | ···················· | ···················· | | |
| B | 8004 hCoV-19/England/CAMB-788FC/2020\|EPI_ISL_433790\|202 | ···················· | ···················· | ···················· | ···················· | ···················· | ···················· | | |
| B | 8005 hCoV-19/England/CAMB-78917/2020\|EPI_ISL_433791\|202 | ···················· | ···················· | ···················· | ···················· | ···················· | ···················· | | |
| B | 8006 hCoV-19/England/CAMB-78926/2020\|EPI_ISL_433792\|202 | ···················· | ···················· | ···················· | ···················· | ···················· | ···················· | | |
| B | 8007 hCoV-19/England/CAMB-78935/2020\|EPI_ISL_433793\|202 | ···················· | ···················· | ···················· | ···················· | ···················· | ···················· | | |
| B | 8008 hCoV-19/England/CAMB-78944/2020\|EPI_ISL_433794\|202 | ···················· | ···················· | ···················· | ···················· | ···················· | ···················· | | |
| B | 8009 hCoV-19/England/CAMB-78953/2020\|EPI_ISL_433795\|202 | ···················· | ···················· | ···················· | ···················· | ···················· | ···················· | | |
| B | 8010 hCoV-19/England/CAMB-78962/2020\|EPI_ISL_433796\|202 | ···················· | ···················· | ···················· | ···················· | ···················· | ···················· | | |
| B | 8011 hCoV-19/England/CAMB-78971/2020\|EPI_ISL_433797\|202 | ···················· | ···················· | ···················· | ···················· | ···················· | ···················· | | |
| B | 8012 hCoV-19/England/CAMB-78980/2020\|EPI_ISL_433798\|202 | ···················· | ···················· | ···················· | ···················· | ···················· | ···················· | | |
| B | 8013 hCoV-19/England/CAMB-7899F/2020\|EPI_ISL_433799\|202 | ···················· | ···················· | ···················· | ···················· | ···················· | ···················· | | |
| B | 8014 hCoV-19/Austria/Graz-MUG4/2020\|EPI_ISL_437200\|2020 | ···················· | ···················· | ···················· | ···················· | ···················· | ···················· | | |
| B | 8015 hCoV-19/Austria/Graz-MUG6/2020\|EPI_ISL_437202\|2020 | ···················· | ···················· | ···················· | ···················· | ···················· | ···················· | | |
| B | 8016 hCoV-19/Austria/Graz-MUG7/2020\|EPI_ISL_437203\|2020 | ···················· | ···················· | ·····T·············· | ···················· | ···················· | ···················· | | |
| B | 8017 hCoV-19/Germany/BY-MVP-0002/2020\|EPI_ISL_437204\|20 | ···················· | ···················· | ···················· | ···················· | ···················· | ···················· | | |
| B | 8018 hCoV-19/Germany/BY-MVP-0003/2020\|EPI_ISL_437205\|20 | ···················· | ···················· | ···················· | ···················· | ···················· | ···················· | | |
| B | 8019 hCoV-19/Germany/BY-MVP-0005/2020\|EPI_ISL_437206\|20 | ···················· | ···················· | ···················· | ···················· | ···················· | ···················· | | |
| B | 8020 hCoV-19/Germany/BY-MVP-0006/2020\|EPI_ISL_437207\|20 | ···················· | ···················· | ···················· | ···················· | ···················· | ···················· | | |
| B | 8021 hCoV-19/Germany/BY-MVP-0051/2020\|EPI_ISL_437250\|20 | ···················· | ···················· | ···················· | ···················· | ···················· | ···················· | | |
| B | 8022 hCoV-19/Germany/BY-MVP-0052/2020\|EPI_ISL_437251\|20 | ···················· | ···················· | ···················· | ···················· | ···················· | ···················· | | |
| B | 8023 hCoV-19/Germany/BY-MVP-0053/2020\|EPI_ISL_437252\|20 | ···················· | ···················· | ···················· | ···················· | ···················· | ···················· | | |
| B | 8024 hCoV-19/Germany/BY-MVP-0054/2020\|EPI_ISL_437253\|20 | ···················· | ···················· | ···················· | ···················· | ···················· | ···················· | | |
| B | 8025 hCoV-19/Germany/BY-MVP-0055/2020\|EPI_ISL_437254\|20 | ···················· | ···················· | ···················· | ···················· | ···················· | ···················· | | |
| B | 8026 hCoV-19/Germany/BY-MVP-0056/2020\|EPI_ISL_437255\|20 | ···················· | ···················· | ···················· | ···················· | ···················· | ···················· | | |
| B | 8027 hCoV-19/Germany/BY-MVP-0057/2020\|EPI_ISL_437256\|20 | ···················· | ···················· | ···················· | ···················· | ···················· | ···················· | | |
| B | 8028 hCoV-19/Germany/BY-MVP-0058/2020\|EPI_ISL_437257\|20 | ···················· | ···················· | ···················· | ···················· | ···················· | ···················· | | |
| B | 8029 hCoV-19/Germany/BY-MVP-0059/2020\|EPI_ISL_437258\|20 | ···················· | ···················· | ···················· | ···················· | ···················· | ···················· | | |
| B | 8030 hCoV-19/Germany/BY-MVP-0060/2020\|EPI_ISL_437259\|20 | ···················· | ···················· | ···················· | ···················· | ···················· | ···················· | | |
| B | 8031 hCoV-19/Germany/BY-MVP-0061/2020\|EPI_ISL_437260\|20 | ···················· | ···················· | ···················· | ···················· | ···················· | ···················· | | |
| B | 8032 hCoV-19/Germany/BY-MVP-0062/2020\|EPI_ISL_437261\|20 | ···················· | ···················· | ···················· | ···················· | ···················· | ···················· | | |
| B | 8033 hCoV-19/Germany/BY-MVP-0063/2020\|EPI_ISL_437262\|20 | ···················· | ···················· | ···················· | ···················· | ···················· | ···················· | | |
| B | 8034 hCoV-19/Germany/BY-MVP-0064/2020\|EPI_ISL_437263\|20 | ···················· | C··················· | ···················· | ···················· | ···················· | ···················· | | |
| B | 8035 hCoV-19/Germany/BY-MVP-0067/2020\|EPI_ISL_437264\|20 | ···················· | ···················· | ···················· | ···················· | ···················· | ···················· | | |
| B | 8037 hCoV-19/Germany/BY-MVP-0070/2020\|EPI_ISL_437266\|20 | ···················· | ···················· | ···················· | ···················· | ···················· | ···················· | | |
| B | 8038 hCoV-19/Germany/BY-MVP-0071/2020\|EPI_ISL_437267\|20 | ···················· | ···················· | ···················· | ···················· | ···················· | ···················· | | |
| B | 8039 hCoV-19/Germany/BY-MVP-0072/2020\|EPI_ISL_437268\|20 | ···················· | ···················· | ···················· | ···················· | ···················· | ···················· | | |
| B | 8040 hCoV-19/Germany/BY-MVP-0073/2020\|EPI_ISL_437269\|20 | ···················· | ···················· | ···················· | ···················· | ···················· | ···················· | | |
| B | 8041 hCoV-19/Germany/BY-MVP-0075/2020\|EPI_ISL_437270\|20 | ···················· | ···················· | ···················· | ···················· | ···················· | ···················· | | |
| B | 8042 hCoV-19/Germany/BY-MVP-0076/2020\|EPI_ISL_437271\|20 | ···················· | ···················· | ···················· | ···················· | ···················· | ···················· | | |
| B | 8043 hCoV-19/Germany/BY-MVP-0077/2020\|EPI_ISL_437272\|20 | ···················· | ···················· | ···················· | ···················· | ···················· | ···················· | | |
| B | 8044 hCoV-19/Germany/BY-MVP-0079/2020\|EPI_ISL_437273\|20 | ···················· | ···················· | ···················· | ···················· | ···················· | ···················· | | |
| B | 8045 hCoV-19/Germany/BY-MVP-0080/2020\|EPI_ISL_437274\|20 | ···················· | ···················· | ···················· | ···················· | ···················· | ··············N···NN | 1 | 1 |
| B | 8046 hCoV-19/Germany/BY-MVP-0081/2020\|EPI_ISL_437275\|20 | ···················· | ···················· | ···················· | ···················· | ···················· | ···················· | | |
| B | 8047 hCoV-19/Germany/BY-MVP-0082/2020\|EPI_ISL_437276\|20 | ···················· | ···················· | ···················· | ···················· | ···················· | ···················· | | |

| | | | | | | | | |
|---|---|---|---|---|---|---|---|---|
| B | 8048 | hCoV-19/Germany/BY-MVP-0083/2020\|EPI_ISL_437277\|20 | ···················· | ···················· | ···················· | ···················· | ···················· | ···················· |
| B | 8049 | hCoV-19/Germany/BY-MVP-0084/2020\|EPI_ISL_437278\|20 | ···················· | ···················· | ···················· | ···················· | ···················· | ···················· |
| B | 8050 | hCoV-19/Germany/BY-MVP-0085/2020\|EPI_ISL_437279\|20 | ···················· | ···················· | ···················· | ···················· | ···················· | ···················· |
| B | 8051 | hCoV-19/Germany/BY-MVP-0086/2020\|EPI_ISL_437280\|20 | ···················· | C··················· | ···················· | ···················· | ···················· | ···················· |
| B | 8052 | hCoV-19/Germany/BY-MVP-0087/2020\|EPI_ISL_437281\|20 | ···················· | ···················· | ···················· | ···················· | ···················· | ···················· |
| B | 8053 | hCoV-19/Germany/BY-MVP-0088/2020\|EPI_ISL_437282\|20 | ···················· | ···················· | ···················· | ···················· | ···················· | ···················· |
| B | 8054 | hCoV-19/Germany/BY-MVP-0089/2020\|EPI_ISL_437283\|20 | ···················· | ···················· | ···················· | ···················· | ···················· | ···················· |
| B | 8055 | hCoV-19/Germany/BY-MVP-0090/2020\|EPI_ISL_437284\|20 | ···················· | ···················· | ···················· | ···················· | ···················· | ···················· |
| B | 8057 | hCoV-19/Germany/BY-MVP-0092/2020\|EPI_ISL_437286\|20 | ···················· | ···················· | ···················· | ···················· | ···················· | ···················· |
| B | 8058 | hCoV-19/Germany/BY-MVP-0095/2020\|EPI_ISL_437287\|20 | ···················· | ···················· | ···················· | ···················· | ···················· | ···················· |
| B | 8059 | hCoV-19/Germany/BY-MVP-0096/2020\|EPI_ISL_437288\|20 | ···················· | ···················· | ···················· | ···················· | ···················· | ···················· |
| B | 8060 | hCoV-19/Germany/BY-MVP-0097/2020\|EPI_ISL_437289\|20 | ···················· | ···················· | ···················· | ···················· | ···················· | ···················· |
| B | 8061 | hCoV-19/Germany/BY-MVP-0099/2020\|EPI_ISL_437290\|20 | ···················· | ···················· | ···················· | ···················· | ···················· | ···················· |
| B | 8062 | hCoV-19/Germany/BY-MVP-0100/2020\|EPI_ISL_437291\|20 | ···················· | ···················· | ···················· | ···················· | ···················· | ···················· |
| B | 8063 | hCoV-19/Germany/BY-MVP-0101/2020\|EPI_ISL_437292\|20 | ···················· | ···················· | ···················· | ···················· | ···················· | ···················· |
| B | 8066 | hCoV-19/Germany/BY-MVP-0106/2020\|EPI_ISL_437296\|20 | ···················· | ···················· | ···················· | ···················· | ···················· | ···················· |
| B | 8067 | hCoV-19/Germany/BY-MVP-0107/2020\|EPI_ISL_437297\|20 | ···················· | ···················· | ···················· | ···················· | ··········N········· | ···················· |
| B | 8068 | hCoV-19/Austria/Graz-MUG8/2020\|EPI_ISL_437298\|2020 | ···················· | ···················· | ···················· | ···················· | ···················· | ···················· |
| B | 8069 | hCoV-19/Austria/Graz-MUG9/2020\|EPI_ISL_437299\|2020 | ···················· | ···················· | ···················· | ···················· | ···················· | ···················· |
| B | 8070 | hCoV-19/England/CAMB-77514/2020\|EPI_ISL_438517\|202 | ···················· | ···················· | ···················· | ···················· | ···················· | ···················· |
| B | 8071 | hCoV-19/England/CAMB-7669A/2020\|EPI_ISL_438518\|202 | ···················· | ···················· | ···················· | ···················· | ···················· | ···················· |
| B | 8072 | hCoV-19/England/CAMB-76E83/2020\|EPI_ISL_438519\|202 | ···················· | ···················· | ···················· | ···················· | ···················· | ···················· |
| B | 8073 | hCoV-19/England/CAMB-77541/2020\|EPI_ISL_438520\|202 | ···················· | ···················· | ···················· | ···················· | ···················· | ···················· |
| B | 8074 | hCoV-19/England/CAMB-763DF/2020\|EPI_ISL_438521\|202 | ···················· | ···················· | ···················· | ···················· | ···················· | ···················· |
| B | 8075 | hCoV-19/England/CAMB-77505/2020\|EPI_ISL_438523\|202 | ···T················ | ···················· | ···················· | ···················· | ···················· | ···················· |
| B | 8076 | hCoV-19/England/CAMB-76DE0/2020\|EPI_ISL_438524\|202 | ···················· | ···················· | ···················· | ···················· | ···················· | ···················· |
| B | 8077 | hCoV-19/England/CAMB-77954/2020\|EPI_ISL_438525\|202 | ···················· | ···················· | ···················· | ···················· | ···················· | ···················· |
| B | 8078 | hCoV-19/England/CAMB-773A1/2020\|EPI_ISL_438527\|202 | ···················· | ···················· | ···················· | ···················· | ···················· | ···················· |
| B | 8079 | hCoV-19/England/CAMB-775BA/2020\|EPI_ISL_438528\|202 | ···················· | ···················· | ···················· | ···················· | ···················· | ···················· |
| B | 8080 | hCoV-19/England/CAMB-7606F/2020\|EPI_ISL_438529\|202 | ···················· | ···················· | ···················· | ···················· | ···················· | ······G············· |
| B | 8081 | hCoV-19/England/CAMB-77620/2020\|EPI_ISL_438531\|202 | ···················· | ···················· | ···················· | ···················· | ···················· | ···················· |
| B | 8082 | hCoV-19/England/CAMB-772C2/2020\|EPI_ISL_438532\|202 | ···················· | ···················· | ···················· | ···················· | ···················· | ···················· |
| B | 8083 | hCoV-19/England/CAMB-779BE/2020\|EPI_ISL_438534\|202 | ···················· | ···················· | ···················· | ···················· | ···················· | ···················· |
| B | 8084 | hCoV-19/England/CAMB-77471/2020\|EPI_ISL_438535\|202 | ···················· | ···················· | ···················· | ···················· | ···················· | ···················· |
| B | 8085 | hCoV-19/England/CAMB-77198/2020\|EPI_ISL_438536\|202 | ···················· | ···················· | ···················· | ···················· | ···················· | ···················· |
| B | 8086 | hCoV-19/England/CAMB-7765D/2020\|EPI_ISL_438537\|202 | ···················· | ···················· | ···················· | ···················· | ···················· | ···················· |
| B | 8087 | hCoV-19/England/CAMB-765BB/2020\|EPI_ISL_438538\|202 | ···················· | ···················· | ···················· | ···················· | ···················· | ···················· |
| B | 8088 | hCoV-19/England/CAMB-811E6/2020\|EPI_ISL_438580\|202 | ···················· | ···················· | ···················· | ···················· | ···················· | ···················· |
| B | 8089 | hCoV-19/England/CAMB-81E1C/2020\|EPI_ISL_438581\|202 | ···················· | ···················· | ···················· | ···················· | ···················· | ···················· |
| B | 8090 | hCoV-19/England/CAMB-81E3A/2020\|EPI_ISL_438583\|202 | ···················· | ···················· | ···················· | ···················· | ···················· | ···················· |
| B | 8091 | hCoV-19/England/CAMB-81E85/2020\|EPI_ISL_438587\|202 | ···················· | ···················· | ···················· | ···················· | ···················· | ···················· |
| B | 8092 | hCoV-19/England/CAMB-81E94/2020\|EPI_ISL_438588\|202 | ···················· | ···················· | ···················· | ···················· | ···················· | ···················· |
| B | 8093 | hCoV-19/England/CAMB-81EB2/2020\|EPI_ISL_438590\|202 | ···················· | ···G················ | ···················· | ···················· | ···················· | ···················· |
| B | 8094 | hCoV-19/England/CAMB-81ED0/2020\|EPI_ISL_438592\|202 | ···················· | ···················· | ···················· | ···················· | ···················· | ···················· |
| B | 8095 | hCoV-19/England/CAMB-81EEF/2020\|EPI_ISL_438593\|202 | ···················· | ···················· | ···················· | ···················· | ···················· | ···················· |
| B | 8096 | hCoV-19/England/CAMB-81F0A/2020\|EPI_ISL_438594\|202 | ···················· | ···················· | ···················· | ···················· | ···················· | ···················· |
| B | 8097 | hCoV-19/England/CAMB-81F46/2020\|EPI_ISL_438597\|202 | ···················· | ···················· | ···················· | ···················· | ···················· | ···················· |
| B | 8098 | hCoV-19/England/CAMB-81F73/2020\|EPI_ISL_438599\|202 | ···················· | ···················· | ···················· | ···················· | ···················· | ···················· |
| B | 8099 | hCoV-19/England/LIVE-9A117/2020\|EPI_ISL_439848\|202 | ···················· | ···················· | ···················· | ···················· | ···················· | ···················· |
| B | 8100 | hCoV-19/England/LIVE-9A126/2020\|EPI_ISL_439849\|202 | ···················· | ···················· | ···················· | ···················· | ···················· | ···················· |
| B | 8101 | hCoV-19/England/LIVE-9A1AE/2020\|EPI_ISL_439855\|202 | ···················· | ···················· | ···················· | ···················· | ···················· | ···················· |
| B | 8102 | hCoV-19/England/LIVE-9A1CC/2020\|EPI_ISL_439857\|202 | ···················· | ···················· | ···················· | ···················· | ···················· | ···················· |
| B | 8103 | hCoV-19/England/CAMB-72977/2020\|EPI_ISL_439863\|202 | ···················· | ···················· | ···················· | ···················· | ···················· | ···················· |
| B | 8104 | hCoV-19/England/CAMB-72AB0/2020\|EPI_ISL_439865\|202 | ···················· | ···················· | ···················· | ···················· | ···················· | ···················· |
| B | 8105 | hCoV-19/England/CAMB-71FD0/2020\|EPI_ISL_439867\|202 | ···················· | ···················· | ···················· | ···················· | ···················· | ···················· |
| B | 8106 | hCoV-19/England/CAMB-72A56/2020\|EPI_ISL_439868\|202 | ···················· | ···················· | ···················· | ···················· | ···················· | ···················· |
| B | 8107 | hCoV-19/England/CAMB-77BC7/2020\|EPI_ISL_440832\|202 | ···················· | ···················· | ···················· | ···················· | ···················· | ···················· |
| B | 8108 | hCoV-19/England/CAMB-77A60/2020\|EPI_ISL_440839\|202 | ···················· | ···················· | ···················· | ···················· | ···················· | ···················· |
| B | 8109 | hCoV-19/England/LIVE-9BA1B/2020\|EPI_ISL_440894\|202 | ···················· | ···················· | ···················· | ···················· | ···················· | ···················· |
| B | 8110 | hCoV-19/England/LIVE-9BA39/2020\|EPI_ISL_440895\|202 | ···················· | ···················· | ···················· | ···················· | ···················· | ···················· |
| B | 8111 | hCoV-19/England/LIVE-9BA84/2020\|EPI_ISL_440897\|202 | ···················· | ···················· | ···················· | ···················· | ···················· | ···················· |
| B | 8112 | hCoV-19/England/LIVE-9BAA2/2020\|EPI_ISL_440898\|202 | ···················· | ···················· | ···················· | ···················· | ···················· | ···················· |
| B | 8113 | hCoV-19/England/CAMB-81C8B/2020\|EPI_ISL_443100\|202 | ···················· | ···················· | ···················· | ···················· | ···················· | ···················· |
| B | 8114 | hCoV-19/England/CAMB-81948/2020\|EPI_ISL_443101\|202 | ···················· | ···················· | ···················· | ···················· | ···················· | ···················· |
| B | 8115 | hCoV-19/England/CAMB-813A4/2020\|EPI_ISL_443102\|202 | ···················· | ···················· | ···················· | ···················· | ···················· | ···················· |
| B | 8116 | hCoV-19/England/CAMB-806E8/2020\|EPI_ISL_443103\|202 | ···················· | ···················· | ···················· | ···················· | ···················· | ···················· |
| B | 8117 | hCoV-19/England/CAMB-81535/2020\|EPI_ISL_443104\|202 | ···················· | ···················· | ···················· | ···················· | ···················· | ···················· |
| B | 8118 | hCoV-19/England/CAMB-8182D/2020\|EPI_ISL_443105\|202 | ···················· | ···················· | ···················· | ···················· | ···················· | ···················· |
| B | 8119 | hCoV-19/England/CAMB-8185A/2020\|EPI_ISL_443111\|202 | ···················· | ···················· | ···················· | ···················· | ···················· | ···················· |
| B | 8120 | hCoV-19/England/CAMB-817E4/2020\|EPI_ISL_443112\|202 | ···················· | ···················· | ···················· | ···················· | ···················· | ···················· |
| B | 8121 | hCoV-19/England/CAMB-81483/2020\|EPI_ISL_443113\|202 | ···················· | ···················· | ···················· | ···················· | ···················· | ···················· |
| B | 8122 | hCoV-19/England/CAMB-7E805/2020\|EPI_ISL_443114\|202 | ···················· | ···················· | ···················· | ···················· | ···················· | ···················· |
| B | 8123 | hCoV-19/England/CAMB-81966/2020\|EPI_ISL_443116\|202 | ···················· | ···················· | ···················· | ···················· | ···················· | ···················· |
| B | 8124 | hCoV-19/England/CAMB-7FCCC/2020\|EPI_ISL_444402\|202 | ···················· | ···················· | ···················· | ···················· | ···················· | ···················· |
| B | 8125 | hCoV-19/England/CAMB-7FD32/2020\|EPI_ISL_444407\|202 | ···················· | ···················· | ···················· | ···················· | ···················· | ···················· |
| B | 8126 | hCoV-19/England/CAMB-7FDAB/2020\|EPI_ISL_444413\|202 | ···················· | ···················· | ···················· | ···················· | ···················· | ···················· |
| B | 8127 | hCoV-19/England/CAMB-7FDC9/2020\|EPI_ISL_444415\|202 | ···················· | ···G················ | ···················· | ···················· | ···················· | ···················· |
| B | 8128 | hCoV-19/England/CAMB-7FDD8/2020\|EPI_ISL_444416\|202 | ···················· | ···················· | ···················· | ···················· | ···················· | ···················· |

| | | | | | | | | |
|---|---|---|---|---|---|---|---|---|
| B | 8129 hCoV-19/England/CAMB-7FDE7/2020\|EPI_ISL_444417\|202 | .................... | .................... | .................... | .................... | .................... | .................... | |
| B | 8130 hCoV-19/England/CAMB-7FDF6/2020\|EPI_ISL_444418\|202 | .................... | ...................A | .................... | .................... | .................... | .................... | 1 |
| B | 8131 hCoV-19/England/CAMB-7FE02/2020\|EPI_ISL_444419\|202 | .................... | .................... | .................... | .................... | .................... | .................... | |
| B | 8132 hCoV-19/England/CAMB-7FE11/2020\|EPI_ISL_444420\|202 | .................... | .................... | .................... | .................... | .................... | .................... | |
| B | 8133 hCoV-19/England/CAMB-7FE20/2020\|EPI_ISL_444421\|202 | .................... | .................... | .................... | .................... | .................... | .................... | |
| B | 8134 hCoV-19/England/CAMB-7FE3F/2020\|EPI_ISL_444422\|202 | .................... | .................... | .................... | .................... | .................... | .................... | |
| B | 8135 hCoV-19/England/CAMB-7FE4E/2020\|EPI_ISL_444423\|202 | .................... | .................... | .................... | .................... | .................... | .................... | |
| B | 8136 hCoV-19/England/CAMB-7FEC6/2020\|EPI_ISL_444430\|202 | .................... | .................... | .................... | .................... | .................... | .................... | |
| B | 8137 hCoV-19/England/CAMB-80AFB/2020\|EPI_ISL_444432\|202 | .................... | .................... | .................... | .................... | .................... | .................... | |
| B | 8138 hCoV-19/England/CAMB-80E4A/2020\|EPI_ISL_444434\|202 | .................... | .................... | .................... | .................... | .................... | .................... | |
| B | 8139 hCoV-19/England/CAMB-80E59/2020\|EPI_ISL_444435\|202 | .................... | .................... | .................... | .................... | .................... | .................... | |
| B | 8140 hCoV-19/England/CAMB-80E68/2020\|EPI_ISL_444436\|202 | .................... | .................... | .................... | .................... | .................... | .................... | |
| B | 8141 hCoV-19/England/CAMB-80E77/2020\|EPI_ISL_444437\|202 | .................... | .................... | .................... | .................... | .................... | .................... | |
| B | 8142 hCoV-19/England/CAMB-80E86/2020\|EPI_ISL_444438\|202 | .................... | .................... | .................... | .................... | .................... | .................... | |
| B | 8143 hCoV-19/England/CAMB-80E95/2020\|EPI_ISL_444439\|202 | .................... | .................... | .................... | .................... | .................... | .................... | |
| B | 8144 hCoV-19/England/CAMB-80EA4/2020\|EPI_ISL_444440\|202 | .................... | .................... | .................... | .................... | .................... | .................... | |
| B | 8145 hCoV-19/England/CAMB-80EB3/2020\|EPI_ISL_444441\|202 | .................... | .................... | .................... | .................... | .................... | .................... | |
| B | 8146 hCoV-19/England/CAMB-80EC2/2020\|EPI_ISL_444442\|202 | .................... | .................... | .................... | .................... | .................... | .................... | |
| B | 8147 hCoV-19/England/CAMB-80ED1/2020\|EPI_ISL_444443\|202 | .................... | .................... | .................... | .................... | .................... | .................... | |
| B | 8148 hCoV-19/England/CAMB-80EFF/2020\|EPI_ISL_444445\|202 | .................... | .................... | .................... | .................... | .................... | .................... | |
| B | 8149 hCoV-19/England/CAMB-80F0B/2020\|EPI_ISL_444446\|202 | .................... | .................... | .................... | .................... | .................... | .................... | |
| B | 8150 hCoV-19/England/CAMB-80F1A/2020\|EPI_ISL_444447\|202 | .................... | .................... | .................... | .................... | .................... | .................... | |
| B | 8151 hCoV-19/England/CAMB-80F29/2020\|EPI_ISL_444448\|202 | .................... | .................... | .................... | .................... | .................... | .................... | |
| B | 8152 hCoV-19/England/CAMB-80F38/2020\|EPI_ISL_444449\|202 | .................... | .................... | .................... | .................... | .................... | .................... | |
| B | 8153 hCoV-19/England/CAMB-80F47/2020\|EPI_ISL_444450\|202 | .................... | .................... | .................... | .................... | .................... | .................... | |
| B | 8154 hCoV-19/England/CAMB-80F56/2020\|EPI_ISL_444451\|202 | .................... | .................... | .................... | .................... | .................... | .................... | |
| B | 8155 hCoV-19/England/CAMB-80F65/2020\|EPI_ISL_444452\|202 | .................... | .................... | .................... | .................... | .................... | .................... | |
| B | 8156 hCoV-19/England/CAMB-80F74/2020\|EPI_ISL_444453\|202 | .................... | .................... | .................... | .................... | .................... | .................... | |
| B | 8157 hCoV-19/USA/CA-QDX-03/2020\|EPI_ISL_444455\|2020-03- | .................... | .................... | .................... | .................... | .................... | .................... | |
| B | 8158 hCoV-19/India/GJ-GBRC25/2020\|EPI_ISL_444456\|2020-0 | .................... | .................... | .................... | .................... | .................... | .................... | |
| B | 8159 hCoV-19/Wales/PHWC-2B3AC/2020\|EPI_ISL_445701\|2020- | .................... | ....A............... | .................... | .................... | .................... | .................... | |
| B | 8160 hCoV-19/Wales/PHWC-2B3BB/2020\|EPI_ISL_445702\|2020- | .................... | .................... | .................... | .................... | .................... | .................... | |
| B | 8161 hCoV-19/Wales/PHWC-2B3CA/2020\|EPI_ISL_445703\|2020- | .................... | .................... | .................... | .................... | .................... | .................... | |
| B | 8162 hCoV-19/Wales/PHWC-2B3E8/2020\|EPI_ISL_445705\|2020- | .................... | .................... | .................... | .................... | .................... | .................... | |
| B | 8163 hCoV-19/Wales/PHWC-2B412/2020\|EPI_ISL_445708\|2020- | .................... | .................... | .................... | .................... | .................... | .................... | |
| B | 8164 hCoV-19/Wales/PHWC-2B421/2020\|EPI_ISL_445709\|2020- | .................... | .................... | .................... | .................... | .................... | .................... | |
| B | 8165 hCoV-19/Wales/PHWC-2B4A9/2020\|EPI_ISL_445716\|2020- | .................... | .................... | .................... | .................... | .................... | .................... | |
| B | 8166 hCoV-19/Wales/PHWC-2B4D6/2020\|EPI_ISL_445719\|2020- | .................... | ....A............... | .................... | .................... | .................... | .................... | |
| B | 8167 hCoV-19/Wales/PHWC-2B4E5/2020\|EPI_ISL_445720\|2020- | .................... | .................... | .................... | .................... | .................... | .................... | |
| B | 8168 hCoV-19/Wales/PHWC-2B500/2020\|EPI_ISL_445721\|2020- | .................... | ....A............... | .................... | .................... | .................... | .................... | |
| B | 8169 hCoV-19/Wales/PHWC-2B5F1/2020\|EPI_ISL_445734\|2020- | .................... | .................... | .................... | .................... | .................... | .................... | |
| B | 8170 hCoV-19/Wales/PHWC-2B60D/2020\|EPI_ISL_445735\|2020- | .................... | .................... | .................... | .................... | .................... | .................... | |
| B | 8171 hCoV-19/Wales/PHWC-2B62B/2020\|EPI_ISL_445737\|2020- | .................... | .................... | .................... | .................... | .................... | .................... | |
| B | 8172 hCoV-19/Wales/PHWC-2B6A3/2020\|EPI_ISL_445745\|2020- | .................... | .................... | .................... | .................... | .................... | .................... | |
| B | 8173 hCoV-19/Wales/PHWC-2B6B2/2020\|EPI_ISL_445746\|2020- | .................... | .................... | .................... | .................... | .................... | .................... | |
| B | 8174 hCoV-19/Wales/PHWC-2B6C1/2020\|EPI_ISL_445747\|2020- | .................... | .................... | .................... | .................... | .................... | .................... | |
| B | 8175 hCoV-19/Wales/PHWC-2B6D0/2020\|EPI_ISL_445748\|2020- | .................... | .................... | .................... | .................... | .................... | .................... | |
| B | 8176 hCoV-19/Wales/PHWC-2B728/2020\|EPI_ISL_445753\|2020- | .................... | .................... | .................... | .................... | .................... | .................... | |
| B | 8177 hCoV-19/Wales/PHWC-2B737/2020\|EPI_ISL_445754\|2020- | .................... | .................... | .................... | .................... | .................... | .................... | |
| B | 8178 hCoV-19/Wales/PHWC-2B746/2020\|EPI_ISL_445755\|2020- | .................... | .................... | .................... | .................... | .................... | .................... | |
| B | 8179 hCoV-19/Wales/PHWC-2B755/2020\|EPI_ISL_445756\|2020- | .................... | ....A............... | .................... | .................... | .................... | .................... | |
| B | 8180 hCoV-19/Wales/PHWC-2B791/2020\|EPI_ISL_445760\|2020- | .................... | .................... | .................... | .................... | .................... | .................... | |
| B | 8181 hCoV-19/Wales/PHWC-2B7A0/2020\|EPI_ISL_445761\|2020- | .................... | .................... | .................... | .................... | .................... | .................... | |
| B | 8182 hCoV-19/Wales/PHWC-2B7BF/2020\|EPI_ISL_445762\|2020- | .................... | .................... | .................... | .................... | .................... | .................... | |
| B | 8183 hCoV-19/Wales/PHWC-2B7CE/2020\|EPI_ISL_445763\|2020- | .................... | .................... | .................... | .................... | .................... | .................... | |
| B | 8184 hCoV-19/Wales/PHWC-2B816/2020\|EPI_ISL_445766\|2020- | .................... | .................... | .................... | .................... | .................... | .................... | |
| B | 8185 hCoV-19/Wales/PHWC-2B825/2020\|EPI_ISL_445767\|2020- | .................... | .A.................. | .................... | .................... | .................... | .................... | |
| B | 8186 hCoV-19/Wales/PHWC-2B843/2020\|EPI_ISL_445769\|2020- | .................... | .................... | .................... | .................... | .................... | .................... | |
| B | 8187 hCoV-19/Wales/PHWC-2B8AD/2020\|EPI_ISL_445775\|2020- | .................... | .................... | .................... | .................... | .................... | .................... | |
| B | 8188 hCoV-19/Wales/PHWC-2B922/2020\|EPI_ISL_445782\|2020- | .................... | .................... | .................... | .................... | .................... | .................... | |
| B | 8189 hCoV-19/Wales/PHWC-2B97D/2020\|EPI_ISL_445786\|2020- | .................... | .................... | .................... | .................... | .................... | .................... | |
| B | 8190 hCoV-19/Australia/VIC182/2020\|EPI_ISL_450212\|2020- | .................... | .................... | .................... | .................... | .................... | .................... | |
| B | 8191 hCoV-19/Australia/VIC349/2020\|EPI_ISL_450213\|2020- | .................... | .................... | .................... | .................... | .................... | .................... | |
| B | 8193 hCoV-19/Wales/PHWC-285A9/2020\|EPI_ISL_432357\|2020- | .................... | .................... | .................... | .................... | .................... | .................... | |
| B | 8194 hCoV-19/Wales/PHWC-2B263/2020\|EPI_ISL_432358\|2020- | .................... | .................... | .................... | .................... | .................... | .................... | |
| B | 8195 hCoV-19/Wales/PHWC-2863D/2020\|EPI_ISL_432359\|2020- | .................... | .................... | .................... | .................... | .................... | .................... | |
| B | 8196 hCoV-19/Wales/PHWC-2849D/2020\|EPI_ISL_432360\|2020- | .................... | .................... | .................... | .................... | .................... | .................... | |
| B | 8197 hCoV-19/Wales/PHWC-274F8/2020\|EPI_ISL_432361\|2020- | .................... | .................... | .................... | .................... | .................... | .................... | |
| B | 8198 hCoV-19/Wales/PHWC-27C3C/2020\|EPI_ISL_432362\|2020- | .................... | .................... | .................... | .................... | .................... | .................... | |
| B | 8199 hCoV-19/Wales/PHWC-28819/2020\|EPI_ISL_432365\|2020- | .................... | .................... | .................... | .................... | .................... | .................... | |
| B | 8200 hCoV-19/Wales/PHWC-28688/2020\|EPI_ISL_432366\|2020- | .................... | .................... | .................... | .................... | .................... | .................... | |
| B | 8201 hCoV-19/Wales/PHWC-27B20/2020\|EPI_ISL_432367\|2020- | .................... | .................... | .................... | .................... | .................... | ............G....... | |
| B | 8202 hCoV-19/Wales/PHWC-27AD8/2020\|EPI_ISL_432369\|2020- | .................... | .................... | .................... | .................... | .................... | .................... | |
| B | 8203 hCoV-19/Wales/PHWC-288FB/2020\|EPI_ISL_432371\|2020- | .................... | .................... | .................... | .................... | .................... | .................... | |
| B | 8204 hCoV-19/Wales/PHWC-27CC3/2020\|EPI_ISL_432374\|2020- | .................... | .................... | .................... | .................... | .................... | .................... | |
| B | 8205 hCoV-19/Wales/PHWC-27E45/2020\|EPI_ISL_432375\|2020 | .................... | .................... | .................... | .................... | .................... | .................... | |
| B | 8206 hCoV-19/Wales/PHWC-24877/2020\|EPI_ISL_432376\|2020- | .................... | .................... | .................... | .................... | .................... | .................... | |
| B | 8207 hCoV-19/Wales/PHWC-27F7F/2020\|EPI_ISL_432377\|2020- | .................... | .................... | .................... | .................... | .................... | .................... | |

| | | | | | | | | |
|---|---|---|---|---|---|---|---|---|
| B | 8208 | hCoV-19/Wales/PHWC-2B1C0/2020\|EPI_ISL_432378\|2020- | ···················· | ···················· | ···················· | ···················· | ···················· | ···················· |
| B | 8209 | hCoV-19/Wales/PHWC-28424/2020\|EPI_ISL_432379\|2020- | ···················· | ···················· | ···················· | ···················· | ···················· | ···················· |
| B | 8210 | hCoV-19/Wales/PHWC-2B245/2020\|EPI_ISL_432382\|2020- | ···················· | ···················· | ···················· | ···················· | ···················· | ···················· |
| B | 8211 | hCoV-19/Wales/PHWC-29C67/2020\|EPI_ISL_432383\|2020- | ···················· | ···················· | ···················· | ···················· | ···················· | ···················· |
| B | 8212 | hCoV-19/Wales/PHWC-285F4/2020\|EPI_ISL_432385\|2020- | ···················· | ···················· | ···················· | ···················· | ···················· | ···················· |
| B | 8213 | hCoV-19/Wales/PHWC-28433/2020\|EPI_ISL_432387\|2020- | ···················· | ···················· | ···················· | ···················· | ···················· | ···················· |
| B | 8214 | hCoV-19/Wales/PHWC-278B0/2020\|EPI_ISL_432389\|2020- | ···················· | ···················· | ···················· | ···················· | ···················· | ···················· |
| B | 8215 | hCoV-19/Wales/PHWC-271A6/2020\|EPI_ISL_432390\|2020- | ···················· | ···················· | ···················· | ···················· | ···················· | ···················· |
| B | 8216 | hCoV-19/Wales/PHWC-288DD/2020\|EPI_ISL_432393\|2020- | ···················· | ···················· | ···················· | ···················· | ···················· | ···················· |
| B | 8217 | hCoV-19/Wales/PHWC-2858B/2020\|EPI_ISL_432395\|2020- | ···················· | ···················· | ···················· | ···················· | ···················· | ···················· |
| B | 8218 | hCoV-19/Wales/PHWC-2856D/2020\|EPI_ISL_432396\|2020- | ···················· | ···················· | ···················· | ···················· | ···················· | ···················· |
| B | 8219 | hCoV-19/Wales/PHWC-2859A/2020\|EPI_ISL_432397\|2020- | ···················· | ···················· | ···················· | ···················· | ···················· | ···················· |
| B | 8220 | hCoV-19/Wales/PHWC-2ADDC/2020\|EPI_ISL_432399\|2020- | ···················· | ···················· | ···················· | ···················· | ···················· | ···················· |
| B | 8221 | hCoV-19/England/CAMB-73D2F/2020\|EPI_ISL_433688\|202 | ···················· | ···················· | ···················· | ···················· | ···················· | ···················· |
| B | 8222 | hCoV-19/England/CAMB-73D3E/2020\|EPI_ISL_433689\|202 | ···················· | ···················· | ···················· | ···················· | ···················· | ···················· |
| B | 8223 | hCoV-19/England/CAMB-73D4D/2020\|EPI_ISL_433690\|202 | ···················· | ···················· | ···················· | ···················· | ···················· | ···················· |
| B | 8224 | hCoV-19/England/CAMB-73D5C/2020\|EPI_ISL_433691\|202 | ···················· | ···················· | ···················· | ···················· | ···················· | ···················· |
| B | 8225 | hCoV-19/England/CAMB-73D7A/2020\|EPI_ISL_433693\|202 | ···················· | ···················· | ···················· | ···················· | ···················· | ···················· |
| B | 8226 | hCoV-19/England/CAMB-73D89/2020\|EPI_ISL_433694\|202 | ···················· | ···················· | ···················· | ···················· | ···················· | ···················· |
| B | 8227 | hCoV-19/England/CAMB-73D98/2020\|EPI_ISL_433695\|202 | ···················· | ···················· | ···················· | ···················· | ···················· | ···················· |
| B | 8228 | hCoV-19/England/CAMB-73DB6/2020\|EPI_ISL_433697\|202 | ···················· | ···················· | ···················· | ···················· | ···················· | ···················· |
| B | 8229 | hCoV-19/England/CAMB-73DC5/2020\|EPI_ISL_433698\|202 | ···················· | ···················· | ···················· | ···················· | ···················· | ···················· |
| B | 8230 | hCoV-19/England/CAMB-73DD4/2020\|EPI_ISL_433699\|202 | ···················· | ···················· | ···················· | ···················· | ···················· | ···················· |
| B | 8231 | hCoV-19/Austria/Graz-MUG5/2020\|EPI_ISL_437201\|2020 | ···················· | ···················· | ···················· | ···················· | ···················· | ···················· |
| B | 8232 | hCoV-19/Germany/BY-MVP-0007/2020\|EPI_ISL_437208\|20 | ···················· | ···················· | ···················· | ···················· | ···················· | ···················· |
| B | 8233 | hCoV-19/Germany/BY-MVP-0008/2020\|EPI_ISL_437209\|20 | ···················· | ···················· | ···················· | ···················· | ···················· | ···················· |
| B | 8234 | hCoV-19/Germany/BY-MVP-0009/2020\|EPI_ISL_437210\|20 | ···················· | ···················· | ···················· | ···················· | ···················· | ···················· |
| B | 8235 | hCoV-19/Germany/BY-MVP-0010/2020\|EPI_ISL_437211\|20 | ···················· | ···················· | ···················· | ···················· | ···················· | ···················· |
| B | 8236 | hCoV-19/Germany/BY-MVP-0011/2020\|EPI_ISL_437212\|20 | ···················· | ···················· | ···················· | ···················· | ···················· | ···················· |
| B | 8237 | hCoV-19/Germany/BY-MVP-0012/2020\|EPI_ISL_437213\|20 | ···················· | ···················· | ···················· | ···················· | ···················· | ···················· |
| B | 8238 | hCoV-19/Germany/BY-MVP-0013/2020\|EPI_ISL_437214\|20 | ···················· | ···················· | ···················· | ···················· | ···················· | ···················· |
| B | 8239 | hCoV-19/Germany/BY-MVP-0014/2020\|EPI_ISL_437215\|20 | ···················· | ···················· | ···················· | ···················· | ···················· | ···················· |
| B | 8240 | hCoV-19/Germany/BY-MVP-0015/2020\|EPI_ISL_437216\|20 | ···················· | ···················· | ···················· | ···················· | ···················· | ···················· |
| B | 8241 | hCoV-19/Germany/BY-MVP-0016/2020\|EPI_ISL_437217\|20 | ···················· | ···················· | ···················· | ···················· | ···················· | ···················· |
| B | 8242 | hCoV-19/Germany/BY-MVP-0017/2020\|EPI_ISL_437218\|20 | ···················· | ···················· | ···················· | ···················· | ···················· | ···················· |
| B | 8243 | hCoV-19/Germany/BY-MVP-0019/2020\|EPI_ISL_437219\|20 | ···················· | ······A············· | ···················· | ···················· | ···················· | ···················· |
| B | 8244 | hCoV-19/Germany/BY-MVP-0020/2020\|EPI_ISL_437220\|20 | ···················· | ···················· | ···················· | ···················· | ···················· | ···················· |
| B | 8245 | hCoV-19/Germany/BY-MVP-0021/2020\|EPI_ISL_437221\|20 | ···················· | ···················· | ···················· | ···················· | ···················· | ···················· |
| B | 8246 | hCoV-19/Germany/BY-MVP-0022/2020\|EPI_ISL_437222\|20 | ···················· | ···················· | ···················· | ···················· | ···················· | ···················· |
| B | 8247 | hCoV-19/Germany/BY-MVP-0023/2020\|EPI_ISL_437223\|20 | ···················· | ···················· | ···················· | ···················· | ···················· | ···················· |
| B | 8248 | hCoV-19/Germany/BY-MVP-0025/2020\|EPI_ISL_437224\|20 | ···················· | ···················· | ···················· | ···················· | ···················· | ···················· |
| B | 8249 | hCoV-19/Germany/BY-MVP-0026/2020\|EPI_ISL_437225\|20 | ···················· | ···················· | ···················· | ···················· | ···················· | ···················· |
| B | 8250 | hCoV-19/Germany/BY-MVP-0028/2020\|EPI_ISL_437227\|20 | ···················· | ···················· | ···················· | ···················· | ···················· | ···················· |
| B | 8251 | hCoV-19/Germany/BY-MVP-0029/2020\|EPI_ISL_437228\|20 | ···················· | ···················· | ···················· | ···················· | ···················· | ···················· |
| B | 8252 | hCoV-19/Germany/BY-MVP-0030/2020\|EPI_ISL_437229\|20 | ···················· | ···················· | ···················· | ···················· | ···················· | ···················· |
| B | 8253 | hCoV-19/Germany/BY-MVP-0031/2020\|EPI_ISL_437230\|20 | ···················· | ···················· | ···················· | ···················· | ···················· | ···················· |
| B | 8254 | hCoV-19/Germany/BY-MVP-0032/2020\|EPI_ISL_437231\|20 | ···················· | ···················· | ···················· | ···················· | ···················· | ···················· |
| B | 8255 | hCoV-19/Germany/BY-MVP-0033/2020\|EPI_ISL_437232\|20 | ···················· | ···················· | ···················· | ···················· | ···················· | ···················· |
| B | 8256 | hCoV-19/Germany/BY-MVP-0034/2020\|EPI_ISL_437233\|20 | ···················· | ···················· | ···················· | ···················· | ···················· | ···················· |
| B | 8257 | hCoV-19/Germany/BY-MVP-0035/2020\|EPI_ISL_437234\|20 | ···················· | ···················· | ···················· | ···················· | ···················· | ···················· |
| B | 8259 | hCoV-19/Germany/BY-MVP-0037/2020\|EPI_ISL_437236\|20 | ···················· | ···················· | ···················· | ···················· | ···················· | ···················· |
| B | 8260 | hCoV-19/Germany/BY-MVP-0038/2020\|EPI_ISL_437237\|20 | ···················· | ···················· | ···················· | ···················· | ···················· | ···················· |
| B | 8261 | hCoV-19/Germany/BY-MVP-0039/2020\|EPI_ISL_437238\|20 | ···················· | ···················· | ···················· | ···················· | ···················· | ···················· |
| B | 8262 | hCoV-19/Germany/BY-MVP-0040/2020\|EPI_ISL_437239\|20 | ···················· | ···················· | ···················· | ···················· | ···················· | ···················· |
| B | 8263 | hCoV-19/Germany/BY-MVP-0042/2020\|EPI_ISL_437241\|20 | ···················· | ···················· | ···················· | ···················· | ···················· | ···················· |
| B | 8265 | hCoV-19/Germany/BY-MVP-0044/2020\|EPI_ISL_437243\|20 | ···················· | ···················· | ···················· | ···················· | ···················· | ···················· |
| B | 8266 | hCoV-19/Germany/BY-MVP-0045/2020\|EPI_ISL_437244\|20 | ···················· | ···················· | ···················· | ···················· | ···················· | ···················· |
| B | 8267 | hCoV-19/Germany/BY-MVP-0046/2020\|EPI_ISL_437245\|20 | ···················· | ···················· | ···················· | ···················· | ···················· | ···················· |
| B | 8268 | hCoV-19/Germany/BY-MVP-0047/2020\|EPI_ISL_437246\|20 | ···················· | ···················· | ···················· | ···················· | ···················· | ···················· |
| B | 8269 | hCoV-19/Germany/BY-MVP-0048/2020\|EPI_ISL_437247\|20 | ···················· | ···················· | ···················· | ···················· | ···················· | ···················· |
| B | 8270 | hCoV-19/Germany/BY-MVP-0049/2020\|EPI_ISL_437248\|20 | ···················· | ···················· | ···················· | ···················· | ···················· | ···················· |
| B | 8271 | hCoV-19/Germany/BY-MVP-0050/2020\|EPI_ISL_437249\|20 | ···················· | ···················· | ···················· | ···················· | ···················· | ···················· |
| B | 8272 | hCoV-19/England/CAMB-71DE5/2020\|EPI_ISL_438500\|202 | ···················· | ···················· | ···················· | ···················· | ···················· | ···················· |
| B | 8273 | hCoV-19/England/CAMB-75A9F/2020\|EPI_ISL_438501\|202 | ···················· | ···················· | ···················· | ···················· | ···················· | ···················· |
| B | 8274 | hCoV-19/England/CAMB-7683A/2020\|EPI_ISL_438502\|202 | ···················· | ···················· | ···················· | ···················· | ···················· | ···················· |
| B | 8275 | hCoV-19/England/CAMB-75A44/2020\|EPI_ISL_438504\|202 | ···················· | ···················· | ···················· | ···················· | ···················· | ···················· |
| B | 8276 | hCoV-19/England/CAMB-75BD8/2020\|EPI_ISL_438505\|202 | ···················· | ···················· | ···················· | ···················· | ···················· | ···················· |
| B | 8277 | hCoV-19/England/CAMB-766E5/2020\|EPI_ISL_438506\|202 | ···················· | ···················· | ···················· | ···················· | ···················· | ···················· |
| B | 8278 | hCoV-19/England/CAMB-759A1/2020\|EPI_ISL_438507\|202 | ···················· | ···················· | ···················· | ···················· | ···················· | ···················· |
| B | 8279 | hCoV-19/England/CAMB-77277/2020\|EPI_ISL_438509\|202 | ···················· | ···················· | ···················· | ···················· | ···················· | ···················· |
| B | 8280 | hCoV-19/England/CAMB-775F6/2020\|EPI_ISL_438510\|202 | ···················· | ···················· | ···················· | ···················· | ···················· | ···················· |
| B | 8281 | hCoV-19/England/CAMB-771C5/2020\|EPI_ISL_438511\|202 | ···················· | ···················· | ···················· | ···················· | ···················· | ···················· |
| B | 8282 | hCoV-19/England/CAMB-766A9/2020\|EPI_ISL_438512\|202 | ···················· | ···················· | ···················· | ···················· | ···················· | ···················· |
| B | 8283 | hCoV-19/England/CAMB-7757E/2020\|EPI_ISL_438513\|202 | ···················· | ···················· | ···················· | ···················· | ···················· | ···················· |
| B | 8284 | hCoV-19/England/CAMB-777F0/2020\|EPI_ISL_438514\|202 | ···················· | ···················· | ···················· | ···················· | ···················· | ···················· |
| B | 8285 | hCoV-19/England/CAMB-7712F/2020\|EPI_ISL_438515\|202 | ···················· | ···················· | ···················· | ···················· | ···················· | ···················· |
| B | 8286 | hCoV-19/England/CAMB-76BF5/2020\|EPI_ISL_438516\|202 | ···················· | ···················· | ···················· | ···················· | ···················· | ···················· |
| B | 8287 | hCoV-19/England/CAMB-775E7/2020\|EPI_ISL_438539\|202 | ···················· | ···················· | ···················· | ···················· | ···················· | ···················· |

| | | | | | | | | | |
|---|---|---|---|---|---|---|---|---|---|
| B | 8288 | hCoV-19/England/CAMB-77532/2020\|EPI_ISL_438540\|202 | ···················· | ···················· | ···················· | ···················· | ···················· | ···················· | |
| B | 8289 | hCoV-19/England/CAMB-77259/2020\|EPI_ISL_438541\|202 | ···················· | ···················· | ···················· | ···················· | ···················· | ···················· | |
| B | 8290 | hCoV-19/England/CAMB-776F3/2020\|EPI_ISL_438543\|202 | ···················· | ···················· | ···················· | ···················· | ···················· | ···················· | |
| B | 8291 | hCoV-19/England/CAMB-77383/2020\|EPI_ISL_438544\|202 | ···················· | ···················· | ···················· | ···················· | ···················· | ···················· | |
| B | 8292 | hCoV-19/England/CAMB-77453/2020\|EPI_ISL_438545\|202 | ···················· | ···················· | ···················· | ···················· | ···················· | ···················· | |
| B | 8293 | hCoV-19/England/CAMB-80F83/2020\|EPI_ISL_438550\|202 | ···················· | ···················· | ···················· | ···················· | ···················· | ···················· | |
| B | 8294 | hCoV-19/England/CAMB-80FB0/2020\|EPI_ISL_438551\|202 | ···················· | ···················· | ···················· | ···················· | ···················· | ···················· | |
| B | 8295 | hCoV-19/England/CAMB-80FFC/2020\|EPI_ISL_438554\|202 | ···················· | ···················· | ···················· | ···················· | ···················· | ···················· | |
| B | 8296 | hCoV-19/England/CAMB-81025/2020\|EPI_ISL_438557\|202 | ···················· | ···················· | ···················· | ···················· | ···················· | ···················· | |
| B | 8297 | hCoV-19/England/CAMB-81034/2020\|EPI_ISL_438558\|202 | ···················· | ···················· | ···················· | ···················· | ···················· | ···················· | |
| B | 8298 | hCoV-19/England/CAMB-81043/2020\|EPI_ISL_438559\|202 | ···················· | ···················· | ···················· | ···················· | ···················· | ···················· | |
| B | 8299 | hCoV-19/England/CAMB-81061/2020\|EPI_ISL_438560\|202 | ···················· | ···················· | ···················· | ···················· | ···················· | ···················· | |
| B | 8300 | hCoV-19/England/CAMB-81070/2020\|EPI_ISL_438561\|202 | ···················· | ···················· | ···················· | ···················· | ···················· | ···················· | |
| B | 8301 | hCoV-19/England/CAMB-8108F/2020\|EPI_ISL_438562\|202 | ···················· | ···················· | ···················· | ···················· | ···················· | ···················· | |
| B | 8302 | hCoV-19/England/CAMB-8109E/2020\|EPI_ISL_438563\|202 | ···················· | ···················· | ···················· | ···················· | ···················· | ···················· | |
| B | 8303 | hCoV-19/England/CAMB-810AD/2020\|EPI_ISL_438564\|202 | ···················· | ···················· | ···················· | ···················· | ···················· | ···················· | |
| B | 8304 | hCoV-19/England/CAMB-810BC/2020\|EPI_ISL_438565\|202 | ···················· | ···················· | ···················· | ···················· | ···················· | ···················· | |
| B | 8305 | hCoV-19/England/CAMB-810CB/2020\|EPI_ISL_438566\|202 | ···················· | ···················· | ···················· | ···················· | ···················· | ···················· | |
| B | 8306 | hCoV-19/England/CAMB-810E9/2020\|EPI_ISL_438567\|202 | ···················· | ···················· | ···················· | ···················· | ···················· | ···················· | |
| B | 8307 | hCoV-19/England/CAMB-81104/2020\|EPI_ISL_438568\|202 | ···················· | ···················· | ···················· | ···················· | ···················· | ···················· | |
| B | 8308 | hCoV-19/England/CAMB-81131/2020\|EPI_ISL_438571\|202 | ···················· | ···················· | ···················· | ···················· | ···················· | ···················· | |
| B | 8309 | hCoV-19/England/CAMB-81140/2020\|EPI_ISL_438572\|202 | ···················· | ···················· | ···················· | ···················· | ···················· | ···················· | |
| B | 8310 | hCoV-19/England/CAMB-8115F/2020\|EPI_ISL_438573\|202 | ···················· | ···················· | ···················· | ···················· | ···················· | ···················· | |
| B | 8311 | hCoV-19/England/CAMB-8117D/2020\|EPI_ISL_438574\|202 | ···················· | ···················· | ···················· | ···················· | ···················· | ···················· | |
| B | 8312 | hCoV-19/England/CAMB-8118C/2020\|EPI_ISL_438575\|202 | ···················· | ···················· | ···················· | ···················· | ···················· | ···················· | |
| B | 8313 | hCoV-19/England/CAMB-8119B/2020\|EPI_ISL_438576\|202 | ···················· | ···················· | ···················· | ···················· | ···················· | ···················· | |
| B | 8314 | hCoV-19/England/CAMB-811AA/2020\|EPI_ISL_438577\|202 | ···················· | ···················· | ···················· | ···················· | ···················· | ···················· | |
| B | 8315 | hCoV-19/England/CAMB-811B9/2020\|EPI_ISL_438578\|202 | ···················· | ···················· | ···················· | ···················· | ···················· | ···················· | |
| B | 8316 | hCoV-19/England/CAMB-811C8/2020\|EPI_ISL_438579\|202 | ···················· | ···················· | ···················· | ···················· | ···················· | ···················· | |
| B | 8317 | hCoV-19/England/LIVE-99EE4/2020\|EPI_ISL_439827\|202 | ···················· | ···················· | ···················· | ···················· | ···················· | ···················· | |
| B | 8318 | hCoV-19/England/LIVE-99F69/2020\|EPI_ISL_439832\|202 | ···················· | ···················· | ···················· | ···················· | ···················· | ···················· | |
| B | 8319 | hCoV-19/England/LIVE-99F96/2020\|EPI_ISL_439834\|202 | ···················· | ···················· | ···················· | ···················· | ···················· | ···················· | |
| B | 8320 | hCoV-19/England/LIVE-99FF0/2020\|EPI_ISL_439836\|202 | ···················· | ···················· | ···················· | ···················· | ···················· | ···················· | |
| B | 8321 | hCoV-19/England/LIVE-9A01A/2020\|EPI_ISL_439837\|202 | ···················· | ···················· | ···················· | ···················· | ···················· | ···················· | |
| B | 8322 | hCoV-19/England/LIVE-9A26F/2020\|EPI_ISL_439861\|202 | ············T······· | ···················· | ···················· | ···················· | ···················· | ···················· | |
| B | 8323 | hCoV-19/England/CAMB-72898/2020\|EPI_ISL_439872\|202 | ···················· | ···················· | ···················· | ···················· | ···················· | ···················· | |
| B | 8324 | hCoV-19/England/CAMB-728C5/2020\|EPI_ISL_439874\|202 | ···················· | ···················· | ···················· | ···················· | ···················· | ···················· | |
| B | 8325 | hCoV-19/England/CAMB-7250A/2020\|EPI_ISL_439875\|202 | ···················· | ···················· | ···················· | ···················· | ···················· | ···················· | |
| B | 8326 | hCoV-19/England/CAMB-71D12/2020\|EPI_ISL_439876\|202 | ···················· | ···················· | ···················· | ···················· | ···················· | ···················· | |
| B | 8327 | hCoV-19/England/CAMB-7283E/2020\|EPI_ISL_439877\|202 | ···················· | ···················· | ···················· | ···················· | ···················· | ···················· | |
| B | 8328 | hCoV-19/England/CAMB-71C42/2020\|EPI_ISL_439880\|202 | ···················· | ···················· | ···················· | ···················· | ···················· | ···················· | |
| B | 8329 | hCoV-19/England/CAMB-72704/2020\|EPI_ISL_439882\|202 | ···················· | ···················· | ···················· | ···················· | ···················· | ···················· | |
| B | 8330 | hCoV-19/England/CAMB-72A0B/2020\|EPI_ISL_439883\|202 | ···················· | ···················· | ···················· | ···················· | ···················· | ···················· | |
| B | 8331 | hCoV-19/England/CAMB-726F8/2020\|EPI_ISL_439884\|202 | ···················· | ···················· | ···················· | ···················· | ···················· | ···················· | |
| B | 8332 | hCoV-19/England/CAMB-7291D/2020\|EPI_ISL_439886\|202 | ···················· | ···················· | ···················· | ···················· | ···················· | ···················· | |
| B | 8333 | hCoV-19/England/CAMB-72995/2020\|EPI_ISL_439888\|202 | ···················· | ···················· | ···················· | ···················· | ···················· | ···················· | |
| B | 8334 | hCoV-19/England/CAMB-72591/2020\|EPI_ISL_439889\|202 | ···················· | ···················· | ···················· | ···················· | ···················· | ···················· | |
| B | 8335 | hCoV-19/England/CAMB-72230/2020\|EPI_ISL_439891\|202 | ···················· | ···················· | ···················· | ···················· | ···················· | ···················· | |
| B | 8336 | hCoV-19/England/CAMB-71B63/2020\|EPI_ISL_439892\|202 | ···················· | ···················· | ···················· | ···················· | ···················· | ···················· | |
| B | 8337 | hCoV-19/England/CAMB-724A3/2020\|EPI_ISL_439895\|202 | ···················· | ····A··············· | ···················· | ···················· | ···················· | ···················· | |
| B | 8338 | hCoV-19/England/CAMB-725EC/2020\|EPI_ISL_439898\|202 | ···················· | ···················· | ···················· | ···················· | ···················· | ···················· | |
| B | 8339 | hCoV-19/England/CAMB-77C2E/2020\|EPI_ISL_440814\|202 | ···················· | ···················· | ···················· | ···················· | ···················· | ···················· | |
| B | 8340 | hCoV-19/England/LIVE-9A29C/2020\|EPI_ISL_440853\|202 | ···················· | ···················· | ···················· | ···················· | ···················· | ···················· | |
| B | 8341 | hCoV-19/England/LIVE-9A40F/2020\|EPI_ISL_440863\|202 | ···················· | ···················· | ···················· | ···················· | ···················· | ···················· | |
| B | 8342 | hCoV-19/England/LIVE-9B626/2020\|EPI_ISL_440864\|202 | ···················· | ···················· | ···················· | ···················· | ···················· | ···················· | |
| B | 8343 | hCoV-19/England/LIVE-9B644/2020\|EPI_ISL_440866\|202 | ···················· | ···················· | ···················· | ···················· | ···················· | ···················· | |
| B | 8344 | hCoV-19/England/LIVE-9B662/2020\|EPI_ISL_440867\|202 | ···················· | ···················· | ···················· | ···················· | ···················· | ···················· | |
| B | 8345 | hCoV-19/England/LIVE-9B723/2020\|EPI_ISL_440873\|202 | ···················· | ···················A | ···················· | ···················· | ···················· | ···················· | 1 |
| B | 8346 | hCoV-19/England/LIVE-9B77E/2020\|EPI_ISL_440875\|202 | ···················· | ···················· | ···················· | ···················· | ···················· | ···················· | |
| B | 8347 | hCoV-19/England/LIVE-9B78D/2020\|EPI_ISL_440876\|202 | ···················· | ···················· | ···················· | ···················· | ···················· | ···················· | |
| B | 8348 | hCoV-19/England/LIVE-9B83F/2020\|EPI_ISL_440878\|202 | ···················· | ·A·················· | ···················· | ···················· | ···················· | ···················· | |
| B | 8349 | hCoV-19/England/LIVE-9B88A/2020\|EPI_ISL_440881\|202 | ···················· | ···················· | ···················· | ···················· | ···················· | ···················· | |
| B | 8350 | hCoV-19/England/LIVE-9B8B7/2020\|EPI_ISL_440883\|202 | ···················· | ·A·················· | ···················· | ···················· | ···················· | ···················· | |
| B | 8351 | hCoV-19/England/LIVE-9B8D5/2020\|EPI_ISL_440884\|202 | ···················· | ···················· | ···················· | ···················· | ···················· | ···················· | |
| B | 8352 | hCoV-19/England/LIVE-9B90F/2020\|EPI_ISL_440886\|202 | ···················· | ···················· | ···················· | ···················· | ···················· | ···················· | |
| B | 8353 | hCoV-19/England/LIVE-9B93C/2020\|EPI_ISL_440887\|202 | ···················· | ···················· | ···················· | ···················· | ···················· | ···················· | |
| B | 8354 | hCoV-19/England/LIVE-9B9C3/2020\|EPI_ISL_440892\|202 | ···················· | ···················· | ···················· | ···················· | ···················· | ···················· | |
| B | 8355 | hCoV-19/England/CAMB-7EF5B/2020\|EPI_ISL_443007\|202 | ···················· | ···················· | ···················· | ···················· | ···················· | ···················· | |
| B | 8356 | hCoV-19/England/CAMB-8131D/2020\|EPI_ISL_443009\|202 | ···················· | ···················· | ···················· | ···················· | ···················· | ···················· | |
| B | 8357 | hCoV-19/England/CAMB-7F190/2020\|EPI_ISL_443010\|202 | ···················· | ···················· | ···················· | ···················· | ···················· | ···················· | |
| B | 8358 | hCoV-19/England/CAMB-80C22/2020\|EPI_ISL_443011\|202 | ···················· | ···················· | ···················· | ···················· | ···················· | ···················· | |
| B | 8359 | hCoV-19/England/CAMB-80BAD/2020\|EPI_ISL_443015\|202 | ···················· | ···················· | ···················· | ···················· | ···················· | ···················· | |
| B | 8360 | hCoV-19/England/CAMB-815DB/2020\|EPI_ISL_443017\|202 | ···················· | ···················· | ···················· | ···················· | ···················· | ···················· | |
| B | 8361 | hCoV-19/England/CAMB-7F00C/2020\|EPI_ISL_443020\|202 | ···················· | ···················· | ···················· | ···················· | ···················· | ···················· | |
| B | 8362 | hCoV-19/England/CAMB-7F7BC/2020\|EPI_ISL_443021\|202 | ···················· | ···················· | ···················· | ···················· | ···················· | ···················· | |
| B | 8363 | hCoV-19/England/CAMB-7F118/2020\|EPI_ISL_443022\|202 | ···················· | ···················· | ···················· | ···················· | ···················· | ···················· | |
| B | 8364 | hCoV-19/England/CAMB-7EF00/2020\|EPI_ISL_443023\|202 | ···················· | ···················· | ···················· | ···················· | ···················· | ···················· | |
| B | 8365 | hCoV-19/England/CAMB-8123E/2020\|EPI_ISL_443024\|202 | ···················· | ···················· | ···················· | ···················· | ···················· | ···················· | |

| | | | | | | | | |
|---|---|---|---|---|---|---|---|---|
| B | 8366 | hCoV-19/England/CAMB-7ECBE/2020\|EPI_ISL_443025\|202 | .................... | .................... | .................... | .................... | .................... | .................... |
| B | 8367 | hCoV-19/England/CAMB-7ECCD/2020\|EPI_ISL_443026\|202 | .................... | .................... | .................... | .................... | .................... | .................... |
| B | 8368 | hCoV-19/England/CAMB-81298/2020\|EPI_ISL_443027\|202 | .................... | .................... | .................... | .................... | .................... | .................... |
| B | 8369 | hCoV-19/England/CAMB-8130E/2020\|EPI_ISL_443029\|202 | .................... | .A.................. | .................... | .................... | .................... | .................... |
| B | 8370 | hCoV-19/England/CAMB-7F1DC/2020\|EPI_ISL_443030\|202 | .................... | .................... | .................... | .................... | .................... | .................... |
| B | 8371 | hCoV-19/England/CAMB-80CC8/2020\|EPI_ISL_443031\|202 | .................... | .................... | .................... | .................... | .................... | .................... |
| B | 8372 | hCoV-19/England/CAMB-7EFF1/2020\|EPI_ISL_443032\|202 | .................... | .................... | .................... | .................... | .................... | .................... |
| B | 8373 | hCoV-19/England/CAMB-815CC/2020\|EPI_ISL_443033\|202 | .................... | .................... | .................... | .................... | .................... | .................... |
| B | 8374 | hCoV-19/England/CAMB-80CAA/2020\|EPI_ISL_443035\|202 | .................... | .................... | .................... | .................... | .................... | .................... |
| B | 8375 | hCoV-19/England/CAMB-80967/2020\|EPI_ISL_443036\|202 | .................... | .................... | .................... | .................... | .................... | .................... |
| B | 8376 | hCoV-19/England/CAMB-7F51C/2020\|EPI_ISL_443037\|202 | .................... | .................... | .................... | .................... | .................... | .................... |
| B | 8377 | hCoV-19/England/CAMB-8124D/2020\|EPI_ISL_443038\|202 | .................... | .................... | .................... | .................... | .................... | .................... |
| B | 8378 | hCoV-19/England/CAMB-7F400/2020\|EPI_ISL_443039\|202 | .................... | .................... | .................... | .................... | .................... | .................... |
| B | 8379 | hCoV-19/England/CAMB-7EC27/2020\|EPI_ISL_443041\|202 | .................... | .................... | .................... | .................... | .................... | .................... |
| B | 8380 | hCoV-19/England/CAMB-80C13/2020\|EPI_ISL_443042\|202 | .................... | .................... | .................... | .................... | .................... | .................... |
| B | 8381 | hCoV-19/England/CAMB-81605/2020\|EPI_ISL_443043\|202 | .................... | .................... | .................... | .................... | .................... | .................... |
| B | 8382 | hCoV-19/England/CAMB-7F35E/2020\|EPI_ISL_443044\|202 | .................... | .................... | .................... | .................... | .................... | .................... |
| B | 8383 | hCoV-19/England/CAMB-80949/2020\|EPI_ISL_443045\|202 | .................... | .................... | .................... | .................... | .................... | .................... |
| B | 8384 | hCoV-19/England/CAMB-80B52/2020\|EPI_ISL_443046\|202 | .................... | .................... | .................... | .................... | .................... | .................... |
| B | 8385 | hCoV-19/England/CAMB-7F4A6/2020\|EPI_ISL_443047\|202 | .................... | .................... | .................... | .................... | .................... | .................... |
| B | 8386 | hCoV-19/England/CAMB-8169C/2020\|EPI_ISL_443090\|202 | .................... | .................... | .................... | .................... | .................... | .................... |
| B | 8387 | hCoV-19/England/CAMB-813D1/2020\|EPI_ISL_443091\|202 | .................... | .................... | .................... | .................... | .................... | .................... |
| B | 8388 | hCoV-19/England/CAMB-7EA69/2020\|EPI_ISL_443092\|202 | .................... | .................... | .................... | .................... | .................... | .................... |
| B | 8389 | hCoV-19/England/CAMB-81AEB/2020\|EPI_ISL_443093\|202 | .................... | .................... | .................... | .................... | .................... | .................... |
| B | 8390 | hCoV-19/England/CAMB-81896/2020\|EPI_ISL_443094\|202 | .................... | .................... | .................... | .................... | .................... | .................... |
| B | 8391 | hCoV-19/England/CAMB-81A54/2020\|EPI_ISL_443095\|202 | .................... | .................... | .................... | .................... | .................... | .................... |
| B | 8392 | hCoV-19/England/CAMB-7E86F/2020\|EPI_ISL_443096\|202 | .................... | .................... | .................... | .................... | .................... | .................... |
| B | 8393 | hCoV-19/England/CAMB-7E7F9/2020\|EPI_ISL_443097\|202 | .................... | .................... | .................... | .................... | .................... | .................... |
| B | 8394 | hCoV-19/England/CAMB-807A9/2020\|EPI_ISL_443098\|202 | .................... | .................... | .................... | .................... | .................... | .................... |
| B | 8395 | hCoV-19/England/CAMB-8175D/2020\|EPI_ISL_443099\|202 | .................... | .................... | .................... | .................... | .................... | .................... |
| B | 8396 | hCoV-19/England/CAMB-1AB84A/2020\|EPI_ISL_444336\|20 | .................... | .................... | .................... | .................... | .................... | .................... |
| B | 8397 | hCoV-19/England/CAMB-1ABE66/2020\|EPI_ISL_444337\|20 | .................... | .................... | .................... | .................... | .................... | .................... |
| B | 8398 | hCoV-19/England/CAMB-1ABE84/2020\|EPI_ISL_444338\|20 | .................... | .................... | .................... | .................... | .................... | .................... |
| B | 8399 | hCoV-19/England/CAMB-1ABE93/2020\|EPI_ISL_444339\|20 | .................... | .................... | .................... | .................... | .................... | .................... |
| B | 8400 | hCoV-19/England/CAMB-1ABEA2/2020\|EPI_ISL_444340\|20 | .................... | .................... | .................... | .................... | .................... | .................... |
| B | 8401 | hCoV-19/England/CAMB-1ABEC0/2020\|EPI_ISL_444342\|20 | .................... | .................... | .................... | .................... | .................... | .................... |
| B | 8402 | hCoV-19/England/CAMB-1ABEDF/2020\|EPI_ISL_444343\|20 | .................... | .................... | .................... | .................... | .................... | .................... |
| B | 8403 | hCoV-19/England/CAMB-1ABEFD/2020\|EPI_ISL_444345\|20 | .................... | .................... | .................... | .................... | .................... | .................... |
| B | 8404 | hCoV-19/England/CAMB-1ABF09/2020\|EPI_ISL_444346\|20 | .................... | .................... | .................... | .................... | .................... | .................... |
| B | 8405 | hCoV-19/England/CAMB-1ABF27/2020\|EPI_ISL_444348\|20 | .................... | .................... | .................... | .................... | .................... | .................... |
| B | 8406 | hCoV-19/England/CAMB-1ABF45/2020\|EPI_ISL_444349\|20 | .................... | .................... | .................... | .................... | .................... | .................... |
| B | 8407 | hCoV-19/England/CAMB-1ABF63/2020\|EPI_ISL_444351\|20 | .................... | .................... | .................... | .................... | .................... | .................... |
| B | 8408 | hCoV-19/England/CAMB-1ABF72/2020\|EPI_ISL_444352\|20 | .................... | .................... | .................... | .................... | .................... | .................... |
| B | 8409 | hCoV-19/England/CAMB-1ABF81/2020\|EPI_ISL_444353\|20 | .................... | .................... | .................... | .................... | .................... | .................... |
| B | 8410 | hCoV-19/England/CAMB-1ABF90/2020\|EPI_ISL_444354\|20 | .................... | .................... | .................... | .................... | .................... | .................... |
| B | 8411 | hCoV-19/England/CAMB-1ABFBE/2020\|EPI_ISL_444355\|20 | .................... | .................... | .................... | .................... | .................... | .................... |
| B | 8412 | hCoV-19/England/CAMB-1ABFCD/2020\|EPI_ISL_444356\|20 | .................... | .................... | .................... | .................... | .................... | .................... |
| B | 8413 | hCoV-19/England/CAMB-1ABFDC/2020\|EPI_ISL_444357\|20 | .................... | .................... | .................... | .................... | .................... | .................... |
| B | 8414 | hCoV-19/England/CAMB-1ABFEB/2020\|EPI_ISL_444358\|20 | .................... | .................... | .................... | .................... | .................... | .................... |
| B | 8415 | hCoV-19/England/CAMB-1AC005/2020\|EPI_ISL_444359\|20 | .................... | .................... | .................... | .................... | .................... | .................... |
| B | 8416 | hCoV-19/England/CAMB-1AC014/2020\|EPI_ISL_444360\|20 | .................... | .................... | .................... | .................... | .................... | .................... |
| B | 8417 | hCoV-19/England/CAMB-1AC023/2020\|EPI_ISL_444361\|20 | .................... | .................... | .................... | .................... | .................... | .................... |
| B | 8418 | hCoV-19/England/CAMB-1AC032/2020\|EPI_ISL_444362\|20 | .................... | .................... | .................... | .................... | .................... | .................... |
| B | 8419 | hCoV-19/England/CAMB-1AC041/2020\|EPI_ISL_444363\|20 | .................... | .................... | .................... | .................... | .................... | .................... |
| B | 8420 | hCoV-19/England/CAMB-1AC050/2020\|EPI_ISL_444364\|20 | .................... | .................... | .................... | .................... | .................... | .................... |
| B | 8421 | hCoV-19/England/CAMB-1AC06F/2020\|EPI_ISL_444365\|20 | .................... | .................... | .................... | .................... | .................... | .................... |
| B | 8422 | hCoV-19/England/CAMB-1AC07E/2020\|EPI_ISL_444366\|20 | .................... | .................... | .................... | .................... | .................... | .................... |
| B | 8423 | hCoV-19/England/CAMB-1AC0AB/2020\|EPI_ISL_444368\|20 | .................... | .................... | .................... | .................... | .................... | .................... |
| B | 8424 | hCoV-19/England/CAMB-1AC0C9/2020\|EPI_ISL_444370\|20 | .................... | .................... | .................... | .................... | .................... | .................... |
| B | 8425 | hCoV-19/England/CAMB-1AC0E7/2020\|EPI_ISL_444371\|20 | .................... | .................... | .................... | .................... | .................... | .................... |
| B | 8426 | hCoV-19/England/CAMB-1AC0F6/2020\|EPI_ISL_444372\|20 | .................... | .................... | .................... | .................... | .................... | .................... |
| B | 8427 | hCoV-19/England/CAMB-1AC120/2020\|EPI_ISL_444374\|20 | .................... | .................... | .................... | .................... | .................... | .................... |
| B | 8428 | hCoV-19/England/CAMB-1AC13F/2020\|EPI_ISL_444375\|20 | .................... | .................... | .................... | .................... | .................... | .................... |
| B | 8429 | hCoV-19/England/CAMB-1AC14E/2020\|EPI_ISL_444376\|20 | .................... | .................... | .................... | .................... | .................... | .................... |
| B | 8430 | hCoV-19/England/CAMB-1AC15D/2020\|EPI_ISL_444377\|20 | .................... | .................... | .................... | .................... | .................... | .................... |
| B | 8431 | hCoV-19/Wales/PHWC-2AB69/2020\|EPI_ISL_445671\|2020- | .................... | ...A................ | .................... | .................... | .................... | .................... |
| B | 8432 | hCoV-19/Wales/PHWC-2AB78/2020\|EPI_ISL_445672\|2020- | .................... | .................... | .................... | .................... | .................... | .................... |
| B | 8433 | hCoV-19/Wales/PHWC-2AB87/2020\|EPI_ISL_445673\|2020- | .................... | .................... | .................... | .................... | .................... | .................... |
| B | 8434 | hCoV-19/Wales/PHWC-2ABA5/2020\|EPI_ISL_445674\|2020- | .................... | .................... | .................... | .................... | .................... | .................... |
| B | 8435 | hCoV-19/Wales/PHWC-2ABB4/2020\|EPI_ISL_445675\|2020- | .................... | .................... | .................... | .................... | .................... | .................... |
| B | 8436 | hCoV-19/Wales/PHWC-2ABD2/2020\|EPI_ISL_445677\|2020- | .................... | .................... | .................... | .................... | .................... | .................... |
| B | 8437 | hCoV-19/Wales/PHWC-2AC0C/2020\|EPI_ISL_445680\|2020- | .................... | .................... | .................... | .................... | .................... | .................... |
| B | 8438 | hCoV-19/Wales/PHWC-2AC1B/2020\|EPI_ISL_445681\|2020- | .................... | .................... | .................... | .................... | .................... | .................... |
| B | 8439 | hCoV-19/Wales/PHWC-2AC39/2020\|EPI_ISL_445682\|2020- | .................... | .................... | .................... | .................... | .................... | .................... |
| B | 8440 | hCoV-19/Wales/PHWC-2AC48/2020\|EPI_ISL_445683\|2020- | .................... | .................... | .................... | .................... | .................... | .................... |
| B | 8441 | hCoV-19/Wales/PHWC-2AC84/2020\|EPI_ISL_445686\|2020- | .................... | .................... | .................... | .................... | .................... | .................... |
| B | 8442 | hCoV-19/Wales/PHWC-2ACA2/2020\|EPI_ISL_445688\|2020- | .................... | .................... | .................... | .................... | .................... | .................... |
| B | 8443 | hCoV-19/Wales/PHWC-2ACB1/2020\|EPI_ISL_445689\|2020- | .................... | .................... | .................... | .................... | .................... | .................... |

| | | | | | | | | | |
|---|---|---|---|---|---|---|---|---|---|
| B | 8444 | hCoV-19/Wales/PHWC-2ACDF/2020\|EPI_ISL_445691\|2020- | .................... | .................... | .................... | .................... | .................... | .................... | |
| B | 8445 | hCoV-19/Wales/PHWC-2ACEE/2020\|EPI_ISL_445692\|2020- | .................... | .................... | .................... | .................... | .................... | .................... | |
| B | 8446 | hCoV-19/Wales/PHWC-2AD27/2020\|EPI_ISL_445696\|2020- | .................... | .................... | .................... | .................... | .................... | .................... | |
| B | 8447 | hCoV-19/Wales/PHWC-2AD45/2020\|EPI_ISL_445697\|2020- | .................... | .................... | .................... | .................... | .................... | .................... | |
| B | 8448 | hCoV-19/Wales/PHWC-2AD54/2020\|EPI_ISL_445698\|2020- | .................... | .................... | .................... | .................... | .................... | .................... | |
| B | 8449 | hCoV-19/USA/CT-Yale-079/2020\|EPI_ISL_431080\|2020-0 | .................... | .................... | .................... | .................... | .................... | .................... | |
| B | 8450 | hCoV-19/USA/CT-Yale-121/2020\|EPI_ISL_431085\|2020-0 | .................... | .................... | .................... | .................... | .................... | .................... | |
| B | 8451 | hCoV-19/Wales/PHWC-28776/2020\|EPI_ISL_432370\|2020- | .................... | .................... | .................... | .................... | .................... | .................... | |
| B | 8452 | hCoV-19/Scotland/EDB1110/2020\|EPI_ISL_433400\|2020- | .................... | ...................A | .................... | .................... | .................... | .................... | 1 |
| B | 8453 | hCoV-19/Scotland/EDB1111/2020\|EPI_ISL_433401\|2020- | .................... | .A.................. | .................... | .................... | .................... | .................... | |
| B | 8454 | hCoV-19/Scotland/EDB1112/2020\|EPI_ISL_433402\|2020- | .................... | .................... | .................... | .................... | .................... | .................... | |
| B | 8455 | hCoV-19/Scotland/EDB1113/2020\|EPI_ISL_433403\|2020- | .................... | .................... | .................... | .................... | .................... | .................... | |
| B | 8456 | hCoV-19/Scotland/EDB1117/2020\|EPI_ISL_433405\|2020- | .................... | .................... | .................... | .................... | .................... | .................... | |
| B | 8457 | hCoV-19/Scotland/EDB1118/2020\|EPI_ISL_433406\|2020- | .................... | .................... | .................... | .................... | .................... | .................... | |
| B | 8458 | hCoV-19/Scotland/EDB112/2020\|EPI_ISL_433407\|2020-0 | .................... | .................... | .................... | .................... | .................... | .................... | |
| B | 8459 | hCoV-19/Scotland/EDB1148/2020\|EPI_ISL_433408\|2020- | .................... | .................... | .................... | .................... | .................... | .................... | |
| B | 8460 | hCoV-19/Scotland/EDB1150/2020\|EPI_ISL_433409\|2020- | .................... | .................... | .................... | .................... | .................... | .................... | |
| B | 8461 | hCoV-19/Scotland/EDB1151/2020\|EPI_ISL_433410\|2020- | .................... | .................... | .................... | .................... | .................... | .................... | |
| B | 8462 | hCoV-19/Scotland/EDB1152/2020\|EPI_ISL_433411\|2020- | .................... | .................... | .................... | .................... | .................... | .................... | |
| B | 8463 | hCoV-19/Scotland/EDB1158/2020\|EPI_ISL_433412\|2020- | .................... | .A.................. | .................... | .................... | .................... | .................... | |
| B | 8464 | hCoV-19/Scotland/EDB1161/2020\|EPI_ISL_433413\|2020- | .................... | .A.................. | .................... | .................... | .................... | .................... | |
| B | 8465 | hCoV-19/Scotland/EDB1163/2020\|EPI_ISL_433414\|2020- | .................... | .................... | .................... | .................... | .................... | .................... | |
| B | 8466 | hCoV-19/Scotland/EDB1164/2020\|EPI_ISL_433415\|2020- | .................... | .A.................. | .................... | .................... | .................... | .................... | |
| B | 8467 | hCoV-19/Scotland/EDB1165/2020\|EPI_ISL_433416\|2020- | .................... | .................... | .................... | .................... | .................... | .................... | |
| B | 8468 | hCoV-19/Scotland/EDB1167/2020\|EPI_ISL_433417\|2020- | .................... | .................... | .................... | .................... | .................... | .................... | |
| B | 8469 | hCoV-19/Scotland/EDB1168/2020\|EPI_ISL_433418\|2020- | .................... | .................... | .................... | .................... | .................... | .................... | |
| B | 8470 | hCoV-19/Scotland/EDB1169/2020\|EPI_ISL_433419\|2020- | .................... | .................... | .................... | .................... | .................... | .................... | |
| B | 8471 | hCoV-19/Scotland/EDB1170/2020\|EPI_ISL_433420\|2020- | .................... | .................... | .................... | .................... | .................... | .................... | |
| B | 8472 | hCoV-19/Scotland/EDB1171/2020\|EPI_ISL_433421\|2020- | .................... | .................... | .................... | .................... | .................... | .................... | |
| B | 8473 | hCoV-19/Scotland/EDB1173/2020\|EPI_ISL_433423\|2020- | .................... | ......A............. | .................... | .................... | .................... | .................... | |
| B | 8474 | hCoV-19/Scotland/EDB1179/2020\|EPI_ISL_433424\|2020- | .................... | .................... | .................... | .................... | .................... | .................... | |
| B | 8475 | hCoV-19/Scotland/EDB1180/2020\|EPI_ISL_433425\|2020- | .................... | .A.................. | .................... | .................... | .................... | .................... | |
| B | 8476 | hCoV-19/Scotland/EDB1182/2020\|EPI_ISL_433426\|2020- | .................... | .................... | .................... | .................... | .................... | .................... | |
| B | 8477 | hCoV-19/Scotland/EDB1197/2020\|EPI_ISL_433428\|2020- | .................... | .................... | .................... | .................... | .................... | .................... | |
| B | 8478 | hCoV-19/Scotland/EDB1198/2020\|EPI_ISL_433429\|2020- | .................... | .................... | .................... | .................... | .................... | .................... | |
| B | 8479 | hCoV-19/Scotland/EDB1207/2020\|EPI_ISL_433430\|2020- | .................... | .................... | .................... | .................... | .................... | .................... | |
| B | 8480 | hCoV-19/Scotland/EDB1216/2020\|EPI_ISL_433431\|2020- | .................... | .................... | .................... | .................... | .................... | .................... | |
| B | 8481 | hCoV-19/Scotland/EDB1220/2020\|EPI_ISL_433432\|2020- | .................... | .................... | .................... | .................... | .................... | .................... | |
| B | 8482 | hCoV-19/Scotland/EDB129/2020\|EPI_ISL_433433\|2020-0 | .................... | .................... | .................... | .................... | .................... | .................... | |
| B | 8483 | hCoV-19/Scotland/EDB1317/2020\|EPI_ISL_433434\|2020- | .................... | .................... | .................... | .................... | .................... | .................... | |
| B | 8484 | hCoV-19/Scotland/EDB1324/2020\|EPI_ISL_433435\|2020- | .................... | .................... | .................... | .................... | .................... | .................... | |
| B | 8485 | hCoV-19/Scotland/EDB1339/2020\|EPI_ISL_433436\|2020- | .................... | .A.................. | .................... | .................... | .................... | .................... | |
| B | 8486 | hCoV-19/Scotland/EDB1340/2020\|EPI_ISL_433437\|2020- | .................... | .................... | .................... | .................... | .................... | .................... | |
| B | 8487 | hCoV-19/Scotland/EDB1404/2020\|EPI_ISL_433439\|2020- | .................... | .................... | .................... | .................... | .................... | .................... | |
| B | 8488 | hCoV-19/Scotland/EDB1411/2020\|EPI_ISL_433440\|2020- | .................... | .A.................. | .................... | .................... | .................... | .................... | |
| B | 8489 | hCoV-19/Scotland/EDB1412/2020\|EPI_ISL_433441\|2020- | .................... | .................... | .................... | .................... | .................... | .................... | |
| B | 8490 | hCoV-19/Scotland/EDB1424/2020\|EPI_ISL_433442\|2020- | .................... | .............A...... | .................... | .................... | .................... | .................... | |
| B | 8491 | hCoV-19/Scotland/EDB1428/2020\|EPI_ISL_433443\|2020- | .................... | .................... | .................... | .................... | .................... | .................... | |
| B | 8492 | hCoV-19/Scotland/EDB1431/2020\|EPI_ISL_433444\|2020- | .................... | .................... | .................... | .................... | .................... | .................... | |
| B | 8493 | hCoV-19/Thailand/NIH-2472/2020\|EPI_ISL_434701\|2020 | .................... | .................... | .................... | .................... | .................... | .................... | |
| B | 8494 | hCoV-19/Thailand/NIH-2489/2020\|EPI_ISL_434703\|2020 | .................... | .................... | .................... | .................... | .................... | .................... | |
| B | 8495 | hCoV-19/Thailand/NIH-2720/2020\|EPI_ISL_434705\|2020 | .................... | .................... | .................... | .................... | .................... | .................... | |
| B | 8496 | hCoV-19/Thailand/NIH-2721/2020\|EPI_ISL_434706\|2020 | .................... | .................... | .................... | .................... | .................... | .................... | |
| B | 8497 | hCoV-19/Thailand/NIH-2982-2/2020\|EPI_ISL_434708\|20 | .................... | .................... | .................... | .................... | .................... | .................... | |
| B | 8499 | hCoV-19/DRC/1423/2020\|EPI_ISL_434710\|2020-04-08 | .................... | .................... | .................... | .................... | .................... | .................... | |
| B | 8500 | hCoV-19/USA/TX-HMH01/2020\|EPI_ISL_434712\|2020-03-1 | .................... | .................... | .................... | .................... | .................... | .................... | |
| B | 8501 | hCoV-19/USA/TX-HMH02/2020\|EPI_ISL_434713\|2020-03-1 | .................... | .................... | .................... | .................... | .................... | .................... | |
| B | 8502 | hCoV-19/USA/TX-HMH03/2020\|EPI_ISL_434714\|2020-03-1 | .................... | .................... | .................... | .................... | .................... | .................... | |
| B | 8503 | hCoV-19/USA/TX-HMH04/2020\|EPI_ISL_434715\|2020-03-1 | .................... | .................... | .................... | .................... | .................... | .................... | |
| B | 8504 | hCoV-19/USA/TX-HMH05/2020\|EPI_ISL_434716\|2020-03-1 | .................... | .................... | .................... | .................... | .................... | .................... | |
| B | 8505 | hCoV-19/USA/TX-HMH06/2020\|EPI_ISL_434717\|2020-03-1 | .................... | .................... | .................... | .................... | .................... | .................... | |
| B | 8506 | hCoV-19/USA/TX-HMH07/2020\|EPI_ISL_434718\|2020-03-1 | .................... | .................... | .................... | .................... | .................... | .................... | |
| B | 8507 | hCoV-19/USA/TX-HMH09/2020\|EPI_ISL_434720\|2020-03-1 | .................... | .................... | .................... | .................... | .................... | .................... | |
| B | 8508 | hCoV-19/USA/TX-HMH010/2020\|EPI_ISL_434721\|2020-03- | .................... | .................... | .................... | .................... | .................... | .................... | |
| B | 8509 | hCoV-19/USA/TX-HMH011/2020\|EPI_ISL_434722\|2020-03- | .................... | .................... | .................... | .................... | .................... | .................... | |
| B | 8510 | hCoV-19/USA/TX-HMH014/2020\|EPI_ISL_434724\|2020-03- | .................... | .................... | .................... | .................... | ..........T......... | .................... | |
| B | 8511 | hCoV-19/USA/TX-HMH015/2020\|EPI_ISL_434725\|2020-03- | .................... | .................... | .................... | .................... | .................... | .................... | |
| B | 8512 | hCoV-19/USA/TX-HMH016/2020\|EPI_ISL_434726\|2020-03- | .................... | .................... | .................... | .................... | .................... | .................... | |
| B | 8513 | hCoV-19/USA/TX-HMH017/2020\|EPI_ISL_434727\|2020-03- | .................... | .................... | .................... | .................... | .................... | .................... | |
| B | 8514 | hCoV-19/USA/TX-HMH018/2020\|EPI_ISL_434728\|2020-03- | .................... | .................... | .................... | .................... | .................... | .................... | |
| B | 8515 | hCoV-19/USA/TX-HMH019/2020\|EPI_ISL_434729\|2020-03- | .................... | .................... | .................... | .................... | .................... | .................... | |
| B | 8516 | hCoV-19/USA/TX-HMH020/2020\|EPI_ISL_434730\|2020-03- | .................... | .................... | .................... | .................... | .................... | .................... | |
| B | 8517 | hCoV-19/USA/TX-HMH021/2020\|EPI_ISL_434731\|2020-03- | .................... | .................... | .................... | .................... | .................... | .................... | |
| B | 8518 | hCoV-19/USA/TX-HMH022/2020\|EPI_ISL_434732\|2020-03- | .................... | .................... | .................... | .................... | .................... | .................... | |
| B | 8520 | hCoV-19/USA/TX-HMH028/2020\|EPI_ISL_434737\|2020-03- | .................... | .................... | .................... | .................... | .................... | .................... | |
| B | 8521 | hCoV-19/USA/TX-HMH035/2020\|EPI_ISL_434744\|2020-03- | .................... | .................... | .................... | .................... | .................... | .................... | |
| B | 8522 | hCoV-19/USA/TX-HMH036/2020\|EPI_ISL_434745\|2020-03- | .................... | .................... | .................... | .................... | .................... | .................... | |
| B | 8524 | hCoV-19/USA/TX-HMH044/2020\|EPI_ISL_434753\|2020-03- | .................... | .................... | .................... | .................... | ..............N..... | NN.................. | |

| | | | | | | | | |
|---|---|---|---|---|---|---|---|---|
| B | 8526 hCoV-19/USA/TX-HMH050/2020\|EPI_ISL_434759\|2020-03- | ···················· | ···················· | ···················· | ···················· | ···················· | ···················· | |
| B | 8528 hCoV-19/USA/TX-HMH055/2020\|EPI_ISL_434764\|2020-03- | ···················· | ···················· | ···················· | ···················· | ···················· | ···················· | |
| B | 8531 hCoV-19/USA/TX-HMH063/2020\|EPI_ISL_434772\|2020-03- | ···················· | ···················· | ···················· | ···················· | ···················· | ···················· | |
| B | 8532 hCoV-19/USA/TX-HMH067/2020\|EPI_ISL_434776\|2020-03- | ···················· | ···················· | ···················· | ···················· | ···················· | ···················· | |
| B | 8533 hCoV-19/USA/WA-S667/2020\|EPI_ISL_438220\|2020-04-21 | ···················· | ···················· | ···················· | ···················· | ···················· | ···················· | |
| B | 8534 hCoV-19/USA/WA-S668/2020\|EPI_ISL_438221\|2020-04-22 | ···················· | ···················· | ···················· | ···················· | ···················· | ···················· | |
| B | 8535 hCoV-19/USA/MD-HP00025/2020\|EPI_ISL_438222\|2020-03 | ···················· | ···················· | ···················· | ···················· | ···················· | ···················· | |
| B | 8536 hCoV-19/USA/DC-HP00026/2020\|EPI_ISL_438223\|2020-03 | ···················· | ···················· | ···················· | ···················· | ···················· | ···················· | |
| B | 8537 hCoV-19/USA/MD-HP00028/2020\|EPI_ISL_438224\|2020-03 | ···················· | ···················· | ···················· | ···················· | ···················· | ···················· | |
| B | 8538 hCoV-19/USA/MD-HP00032/2020\|EPI_ISL_438225\|2020-03 | ···················· | ···················· | ···················· | ···················· | ···················· | ···················· | |
| B | 8539 hCoV-19/USA/MD-HP00033/2020\|EPI_ISL_438226\|2020-03 | ···················· | ···················· | ···················· | ···················· | ···················· | ···················· | |
| B | 8540 hCoV-19/USA/MD-HP00069/2020\|EPI_ISL_438228\|2020-03 | ···················· | ···················· | ···················· | ···················· | ···················· | ···················· | |
| B | 8541 hCoV-19/USA/DC-HP00070/2020\|EPI_ISL_438229\|2020-03 | ···················· | ···················· | ···················· | ···················· | ···················· | ···················· | |
| B | 8542 hCoV-19/USA/MD-HP00075/2020\|EPI_ISL_438233\|2020-03 | ···················· | ···················· | ···················· | ···················· | ···················· | ···················· | |
| B | 8543 hCoV-19/USA/MD-HP00076/2020\|EPI_ISL_438234\|2020-03 | ···················· | ···················· | ···················· | ···················· | ···················· | ···················· | |
| B | 8544 hCoV-19/USA/MD-HP00078/2020\|EPI_ISL_438235\|2020-03 | ···················· | ···················· | ···················· | ···················· | ···················· | ···················· | |
| B | 8545 hCoV-19/USA/DC-HP00080/2020\|EPI_ISL_438237\|2020-03 | ···················· | ···················· | ···················· | ···················· | ···················· | ···················· | |
| B | 8546 hCoV-19/USA/MD-HP00085/2020\|EPI_ISL_438240\|2020-03 | ···················· | ···················· | ···················· | ···················· | ···················· | ···················· | |
| B | 8552 hCoV-19/USA/MD-HP00104/2020\|EPI_ISL_438246\|2020-03 | ···················· | ···················· | ···················· | ···················· | ···················· | ···················· | |
| B | 8554 hCoV-19/England/CAMB-76C6B/2020\|EPI_ISL_438257\|202 | ···················· | ···················· | ···················· | ···················· | ···················· | ···················· | |
| B | 8555 hCoV-19/England/CAMB-7674C/2020\|EPI_ISL_438259\|202 | ···················· | ···················· | ···················· | ···················· | ···················· | ···················· | |
| B | 8556 hCoV-19/England/CAMB-71EB5/2020\|EPI_ISL_439550\|202 | ···················· | ···················· | ···················· | ···················· | ···················· | ···················· | |
| B | 8557 hCoV-19/England/CAMB-71E97/2020\|EPI_ISL_439555\|202 | ···················· | ···················· | ···················· | ···················· | ···················· | ···················· | |
| B | 8558 hCoV-19/England/CAMB-722C7/2020\|EPI_ISL_439557\|202 | ···················· | ···················· | ···················· | ···················· | ···················· | ···················· | |
| B | 8559 hCoV-19/England/CAMB-721AC/2020\|EPI_ISL_439559\|202 | ···················· | ···················· | ···················· | ···················· | ···················· | ···················· | |
| B | 8560 hCoV-19/England/CAMB-7231F/2020\|EPI_ISL_439564\|202 | ···················· | ···················· | ···················· | ···················· | ···················· | ···················· | |
| B | 8561 hCoV-19/England/CAMB-7278C/2020\|EPI_ISL_439565\|202 | ···················· | ···················· | ···················· | ···················· | ···················· | ···················· | |
| B | 8562 hCoV-19/England/CAMB-71C8E/2020\|EPI_ISL_439567\|202 | ···················· | ···················· | ···················· | ···················· | ···················· | ···················· | |
| B | 8563 hCoV-19/England/CAMB-729A4/2020\|EPI_ISL_439568\|202 | ···················· | ···················· | ···················· | ···················· | ···················· | ···················· | |
| B | 8564 hCoV-19/England/CAMB-72670/2020\|EPI_ISL_439570\|202 | ···················· | ···················· | ···················· | ···················· | ···················· | ···················· | |
| B | 8565 hCoV-19/England/CAMB-71D8B/2020\|EPI_ISL_439571\|202 | ···················· | ···················· | ···················· | ···················· | ···················· | ···················· | |
| B | 8566 hCoV-19/England/CAMB-724C1/2020\|EPI_ISL_439572\|202 | ···················· | ···················· | ···················· | ···················· | ···················· | ···················· | |
| B | 8567 hCoV-19/England/CAMB-726AD/2020\|EPI_ISL_439574\|202 | ···················· | ···················· | ···················· | ···················· | ···················· | ···················· | |
| B | 8568 hCoV-19/England/CAMB-7234C/2020\|EPI_ISL_439575\|202 | ···················· | ·A·················· | ···················· | ···················· | ···················· | ···················· | |
| B | 8569 hCoV-19/England/CAMB-72115/2020\|EPI_ISL_439576\|202 | ···················· | ···················· | ···················· | ···················· | ···················· | ···················· | |
| B | 8570 hCoV-19/England/CAMB-72036/2020\|EPI_ISL_439582\|202 | ···················· | ···················· | ···················· | ···················· | ···················· | ···················· | |
| B | 8571 hCoV-19/England/CAMB-720BE/2020\|EPI_ISL_439585\|202 | ···················· | ···················· | ···················· | ···················· | ···················· | ···················· | |
| B | 8572 hCoV-19/England/CAMB-720AF/2020\|EPI_ISL_439587\|202 | ···················· | ···················· | ···················· | ···················· | ···················· | ···················· | |
| B | 8573 hCoV-19/England/CAMB-729C2/2020\|EPI_ISL_439588\|202 | ···················· | ···················· | ···················· | ···················· | ···················· | ···················· | |
| B | 8574 hCoV-19/England/CAMB-71ED3/2020\|EPI_ISL_439589\|202 | ···················· | ···················· | ···················· | ···················· | ···················· | ···················· | |
| B | 8575 hCoV-19/England/LIVE-99BA1/2020\|EPI_ISL_439804\|202 | ···················· | ···················A | ···················· | ···················· | ···················· | ···················· | 1 |
| B | 8576 hCoV-19/England/LIVE-99BCF/2020\|EPI_ISL_439806\|202 | ···················· | ···················· | ···················· | ···················· | ···················· | ···················· | |
| B | 8577 hCoV-19/England/LIVE-99BED/2020\|EPI_ISL_439807\|202 | ···················· | ···················· | ···················· | ···················· | ···················· | ···················· | |
| B | 8578 hCoV-19/England/LIVE-99C08/2020\|EPI_ISL_439808\|202 | ···················· | ·A·················· | ···················· | ···················· | ···················· | ···················· | |
| B | 8579 hCoV-19/England/LIVE-99C44/2020\|EPI_ISL_439810\|202 | ···················· | ···················· | ···················· | ···················· | ···················· | ···················· | |
| B | 8580 hCoV-19/England/LIVE-99CCC/2020\|EPI_ISL_439812\|202 | ···················· | ···················· | ···················· | ···················· | ···················· | ···················· | |
| B | 8581 hCoV-19/England/LIVE-99D14/2020\|EPI_ISL_439815\|202 | ···················· | ···················· | ···················· | ···················· | ···················· | ···················· | |
| B | 8582 hCoV-19/England/LIVE-99D7E/2020\|EPI_ISL_439817\|202 | ···················· | ···················· | ···················· | ···················· | ···················· | ···················· | |
| B | 8583 hCoV-19/England/LIVE-99DD8/2020\|EPI_ISL_439820\|202 | ···················· | ···················· | ···················· | ···················· | ···················· | ···················· | |
| B | 8584 hCoV-19/England/LIVE-99DE7/2020\|EPI_ISL_439821\|202 | ···················· | ···················· | ···················· | ···················· | ···················· | ···················· | |
| B | 8585 hCoV-19/England/LIVE-99EB7/2020\|EPI_ISL_439824\|202 | ···················· | ···················· | ···················· | ···················· | ···················· | ···················· | |
| B | 8586 hCoV-19/England/LIVE-99EC6/2020\|EPI_ISL_439825\|202 | ···················· | ···················· | ···················· | ···················· | ···················· | ···················· | |
| B | 8587 hCoV-19/England/CAMB-848C0/2020\|EPI_ISL_440532\|202 | ···················· | ···················· | ···················· | ···················· | ···················· | ···················· | |
| B | 8588 hCoV-19/England/CAMB-84A42/2020\|EPI_ISL_440533\|202 | ···················· | ···················· | ···················· | ···················· | ···················· | ···················· | |
| B | 8589 hCoV-19/England/CAMB-846E4/2020\|EPI_ISL_440534\|202 | ···················· | ···················· | ···················· | ···················· | ···················· | ···················· | |
| B | 8590 hCoV-19/England/CAMB-846C6/2020\|EPI_ISL_440538\|202 | ···················· | ···················· | ···················· | ···················· | ···················· | ···················· | |
| B | 8591 hCoV-19/England/CAMB-84C1F/2020\|EPI_ISL_440539\|202 | ···················· | ···················· | ···················· | ···················· | ···················· | ···················· | |
| B | 8592 hCoV-19/England/CAMB-84B4F/2020\|EPI_ISL_440541\|202 | ···················· | ···················· | ···················· | ···················· | ···················· | ···················· | |
| B | 8593 hCoV-19/England/CAMB-84C5B/2020\|EPI_ISL_440542\|202 | ···················· | ······A············· | ···················· | ···················· | ···················· | ···················· | |
| B | 8594 hCoV-19/England/CAMB-84B5E/2020\|EPI_ISL_440551\|202 | ···················· | ···················· | ···················· | ···················· | ···················· | ···················· | |
| B | 8595 hCoV-19/England/CAMB-84B03/2020\|EPI_ISL_440552\|202 | ···················· | ···················· | ···················· | ···················· | ···················· | ···················· | |
| B | 8596 hCoV-19/England/CAMB-84B7C/2020\|EPI_ISL_440553\|202 | ···················· | ···················· | ···················· | ···················· | ···················· | ···················· | |
| B | 8597 hCoV-19/England/CAMB-7A6F0/2020\|EPI_ISL_440555\|202 | ···················· | ···················· | ···················· | ···················· | ···················· | ···················· | |
| B | 8598 hCoV-19/England/CAMB-7A13B/2020\|EPI_ISL_440557\|202 | ···················· | ···················· | ···················· | ···················· | ···················· | ···················· | |
| B | 8599 hCoV-19/England/CAMB-79F8D/2020\|EPI_ISL_440558\|202 | ···················· | ···················· | ···················· | ···················· | ···················· | ···················· | |
| B | 8600 hCoV-19/England/CAMB-79B3E/2020\|EPI_ISL_440560\|202 | ···················· | ···················· | ···················· | ···················· | ···················· | ···················· | |
| B | 8601 hCoV-19/England/CAMB-7A3AE/2020\|EPI_ISL_440561\|202 | ···················· | ···················· | ···················· | ···················· | ···················· | ···················· | |
| B | 8602 hCoV-19/England/CAMB-78D2A/2020\|EPI_ISL_440562\|202 | ···················· | ···················· | ···················· | ···················· | ···················· | ···················· | |
| B | 8603 hCoV-19/England/CAMB-7A71B/2020\|EPI_ISL_440563\|202 | ···················· | ···················· | ···················· | ···················· | ···················· | ···················· | |
| B | 8604 hCoV-19/England/CAMB-79BB6/2020\|EPI_ISL_440564\|202 | ···················· | ···················· | ···················· | ···················· | ···················· | ···················· | |
| B | 8605 hCoV-19/England/CAMB-7A9E8/2020\|EPI_ISL_440565\|202 | ···················· | ···················· | ···················· | ···················· | ···················· | ···················· | |
| B | 8606 hCoV-19/England/CAMB-7A3EA/2020\|EPI_ISL_440566\|202 | ···················· | ···················· | ···················· | ···················· | ···················· | ···················· | |
| B | 8607 hCoV-19/England/CAMB-79F41/2020\|EPI_ISL_440567\|202 | ···················· | ···················· | ···················· | ···················· | ···················· | ···················· | |
| B | 8608 hCoV-19/England/CAMB-79FE7/2020\|EPI_ISL_440568\|202 | ···················· | ···················· | ···················· | ···················· | ···················· | ···················· | |
| B | 8609 hCoV-19/England/CAMB-7A784/2020\|EPI_ISL_440569\|202 | ···················· | ···················· | ···················· | ···················· | ···················· | ···················· | |
| B | 8610 hCoV-19/England/CAMB-7A7C0/2020\|EPI_ISL_440570\|202 | ···················· | ···················· | ···················· | ···················· | ···················· | ···················· | |
| B | 8611 hCoV-19/England/CAMB-7A4AB/2020\|EPI_ISL_440571\|202 | ···················· | ···················· | ···················· | ···················· | ···················· | ···················· | |
| B | 8612 hCoV-19/England/CAMB-7A001/2020\|EPI_ISL_440573\|202 | ···················· | ···················· | ···················· | ···················· | ···················· | ···················· | |

| | | | | | | | | |
|---|---|---|---|---|---|---|---|---|
| B | 8613 | hCoV-19/England/BRIS-122975/2020\|EPI_ISL_440800\|20 | ···················· | ···················· | ···················· | ···················· | ···················· | ···················· |
| B | 8614 | hCoV-19/England/BRIS-122A18/2020\|EPI_ISL_440801\|20 | ···················· | ···················· | ···················· | ···················· | ···················· | ···················· |
| B | 8615 | hCoV-19/England/BRIS-1223FF/2020\|EPI_ISL_440802\|20 | ···················· | ···················· | ···················· | ···················· | ···················· | ···················· |
| B | 8616 | hCoV-19/England/BRIS-1229FD/2020\|EPI_ISL_440803\|20 | ···················· | ···················· | ···················· | ···················· | ···················· | ···················· |
| B | 8617 | hCoV-19/England/BRIS-1228C3/2020\|EPI_ISL_440804\|20 | ···················· | ···················· | ···················· | ···················· | ···················· | ···················· |
| B | 8618 | hCoV-19/England/BRIS-122D88/2020\|EPI_ISL_440806\|20 | ···················· | ···················· | ···················· | ···················· | ···················· | ···················· |
| B | 8619 | hCoV-19/England/BRIS-122984/2020\|EPI_ISL_440807\|20 | ···················· | ···················· | ···················· | ···················· | ···················· | ···················· |
| B | 8620 | hCoV-19/England/BRIS-1228E1/2020\|EPI_ISL_440808\|20 | ···················· | ···················· | ···················· | ···················· | ···················· | ···················· |
| B | 8621 | hCoV-19/England/BRIS-122F64/2020\|EPI_ISL_440809\|20 | ···················· | ···················· | ···················· | ···················· | ···················· | ···················· |
| B | 8622 | hCoV-19/England/NOTT-110813/2020\|EPI_ISL_441863\|20 | ···················· | ···················· | ···················· | ···················· | ···················· | ···················· |
| B | 8623 | hCoV-19/England/NOTT-110822/2020\|EPI_ISL_441864\|20 | ···················· | ···················· | ···················· | ···················· | ···················· | ···················· |
| B | 8624 | hCoV-19/England/NOTT-110831/2020\|EPI_ISL_441865\|20 | ···················· | ···················· | ···················· | ···················· | ···················· | ···················· |
| B | 8625 | hCoV-19/England/NOTT-110840/2020\|EPI_ISL_441866\|20 | ···················· | ···················· | ···················· | ···················· | ···················· | ···················· |
| B | 8626 | hCoV-19/England/NOTT-11085F/2020\|EPI_ISL_441867\|20 | ···················· | ···················· | ···················· | ···················· | ···················· | ···················· |
| B | 8627 | hCoV-19/England/NOTT-11086E/2020\|EPI_ISL_441868\|20 | ···················· | ···················· | ···················· | ···················· | ···················· | ···················· |
| B | 8628 | hCoV-19/England/NOTT-11087D/2020\|EPI_ISL_441869\|20 | ···················· | ···················· | ···················· | ···················· | ···················· | ···················· |
| B | 8629 | hCoV-19/England/NOTT-11088C/2020\|EPI_ISL_441870\|20 | ···················· | ···················· | ···················· | ···················· | ···················· | ···················· |
| B | 8630 | hCoV-19/England/NOTT-11089B/2020\|EPI_ISL_441871\|20 | ···················· | ···················· | ···················· | ···················· | ···················· | ···················· |
| B | 8631 | hCoV-19/England/NOTT-1109A7/2020\|EPI_ISL_441880\|20 | ···················· | ···················· | ···················· | ···················· | ···················· | ···················· |
| B | 8632 | hCoV-19/England/NOTT-1109B6/2020\|EPI_ISL_441881\|20 | ···················· | ···················· | ···················· | ···················· | ···················· | ···················· |
| B | 8633 | hCoV-19/England/NOTT-1109C5/2020\|EPI_ISL_441882\|20 | ···················· | ···················· | ···················· | ···················· | ···················· | ···················· |
| B | 8634 | hCoV-19/England/NOTT-1109D4/2020\|EPI_ISL_441883\|20 | ···················· | ···················· | ···················· | ···················· | ···················· | ···················· |
| B | 8635 | hCoV-19/England/NOTT-110A0E/2020\|EPI_ISL_441885\|20 | ···················· | ···················· | ···················· | ···················· | ···················· | ···················· |
| B | 8636 | hCoV-19/England/NOTT-110A1D/2020\|EPI_ISL_441886\|20 | ···················· | ···················· | ···················· | ···················· | ···················· | ···················· |
| B | 8637 | hCoV-19/England/NOTT-110A3B/2020\|EPI_ISL_441888\|20 | ···················· | ···················· | ···················· | ···················· | ···················· | ···················· |
| B | 8638 | hCoV-19/England/NOTT-110A77/2020\|EPI_ISL_441890\|20 | ···················· | ···················· | ···················· | ···················· | ···················· | ···················· |
| B | 8639 | hCoV-19/England/NOTT-110A86/2020\|EPI_ISL_441891\|20 | ···················· | ···················· | ···················· | ···················· | ···················· | ···················· |
| B | 8640 | hCoV-19/England/NOTT-110A95/2020\|EPI_ISL_441892\|20 | ···················· | ···················· | ···················· | ···················· | ···················· | ···················· |
| B | 8641 | hCoV-19/England/NOTT-110AB3/2020\|EPI_ISL_441894\|20 | ···················· | ···················· | ···················· | ···················· | ···················· | ···················· |
| B | 8642 | hCoV-19/England/NOTT-110AC2/2020\|EPI_ISL_441895\|20 | ···················· | ···················· | ···················· | ···················· | ···················· | ···················· |
| B | 8643 | hCoV-19/England/NOTT-110AD1/2020\|EPI_ISL_441896\|20 | ···················· | ···················· | ···················· | ···················· | ···················· | ···················· |
| B | 8644 | hCoV-19/England/NOTT-110AE0/2020\|EPI_ISL_441897\|20 | ···················· | ···················· | ···················· | ···················· | ···················· | ···················· |
| B | 8645 | hCoV-19/England/SHEF-C095C/2020\|EPI_ISL_441898\|202 | ···················· | ···················· | ···················· | ···················· | ···················· | ···················· |
| B | 8646 | hCoV-19/England/CAMB-7F770/2020\|EPI_ISL_443049\|202 | ···················· | ·A·················· | ···················· | ···················· | ···················· | ···················· |
| B | 8647 | hCoV-19/England/CAMB-7EC36/2020\|EPI_ISL_443050\|202 | ···················· | ···················· | ···················· | ···················· | ···················· | ···················· |
| B | 8648 | hCoV-19/England/CAMB-81BE8/2020\|EPI_ISL_443051\|202 | ···················· | ···················· | ···················· | ···················· | ···················· | ···················· |
| B | 8649 | hCoV-19/England/CAMB-7E9E4/2020\|EPI_ISL_443053\|202 | ···················· | ···················· | ···················· | ···················· | ···················· | ···················· |
| B | 8650 | hCoV-19/England/CAMB-81B42/2020\|EPI_ISL_443055\|202 | ···················· | ···················· | ···················· | ···················· | ···················· | ···················· |
| B | 8651 | hCoV-19/England/CAMB-81A18/2020\|EPI_ISL_443058\|202 | ···················· | ···················· | ···················· | ···················· | ···················· | ···················· |
| B | 8652 | hCoV-19/England/CAMB-7E8BA/2020\|EPI_ISL_443059\|202 | ···················· | ···················· | ···················· | ···················· | ···················· | ···················· |
| B | 8653 | hCoV-19/England/CAMB-81C30/2020\|EPI_ISL_443060\|202 | ···················· | ···················· | ···················· | ···················· | ···················· | ···················· |
| B | 8654 | hCoV-19/England/CAMB-7E9B7/2020\|EPI_ISL_443061\|202 | ···················· | ···················· | ···················· | ···················· | ···················· | ···················· |
| B | 8655 | hCoV-19/England/CAMB-8168D/2020\|EPI_ISL_443062\|202 | ···················· | ···················· | ···················· | ···················· | ···················· | ···················· |
| B | 8656 | hCoV-19/England/CAMB-7E999/2020\|EPI_ISL_443063\|202 | ···················· | ···················· | ···················· | ···················· | ···················· | ···················· |
| B | 8657 | hCoV-19/England/CAMB-81447/2020\|EPI_ISL_443065\|202 | ···················· | ···················· | ···················· | ···················· | ···················· | ···················· |
| B | 8658 | hCoV-19/England/CAMB-81939/2020\|EPI_ISL_443066\|202 | ···················· | ···················· | ···················· | ···················· | ···················· | ···················· |
| B | 8659 | hCoV-19/England/CAMB-814DE/2020\|EPI_ISL_443068\|202 | ···················· | ···················· | ···················· | ···················· | ···················· | ···················· |
| B | 8660 | hCoV-19/England/CAMB-81BD9/2020\|EPI_ISL_443070\|202 | ···················· | ···················· | ···················· | ···················· | ···················· | ···················· |
| B | 8661 | hCoV-19/England/CAMB-7E920/2020\|EPI_ISL_443072\|202 | ···················· | ···················· | ···················· | ···················· | ···················· | ···················· |
| B | 8662 | hCoV-19/England/CAMB-7E94E/2020\|EPI_ISL_443074\|202 | ···················· | ···················· | ···················· | ···················· | ···················· | ···················· |
| B | 8663 | hCoV-19/England/CAMB-81DC4/2020\|EPI_ISL_443075\|202 | ···················· | ···················· | ···················· | ···················· | ···················· | ···················· |
| B | 8664 | hCoV-19/England/CAMB-8177B/2020\|EPI_ISL_443076\|202 | ···················· | ···················· | ···················· | ···················· | ···················· | ···················· |
| B | 8665 | hCoV-19/England/CAMB-7E911/2020\|EPI_ISL_443077\|202 | ···················· | ···················· | ···················· | ···················· | ···················· | ···················· |
| B | 8666 | hCoV-19/England/CAMB-818A5/2020\|EPI_ISL_443078\|202 | ···················· | ···················· | ···················· | ···················· | ···················· | ···················· |
| B | 8667 | hCoV-19/England/CAMB-8192A/2020\|EPI_ISL_443080\|202 | ···················· | ···················· | ···················· | ···················· | ···················· | ···················· |
| B | 8668 | hCoV-19/England/CAMB-7E841/2020\|EPI_ISL_443081\|202 | ···················· | ···················· | ···················· | ···················· | ···················· | ···················· |
| B | 8669 | hCoV-19/England/CAMB-81BCA/2020\|EPI_ISL_443083\|202 | ···················· | ···················· | ···················· | ···················· | ···················· | ···················· |
| B | 8670 | hCoV-19/England/CAMB-81A81/2020\|EPI_ISL_443084\|202 | ···················· | ···················· | ···················· | ···················· | ···················· | ···················· |
| B | 8671 | hCoV-19/England/CAMB-81492/2020\|EPI_ISL_443086\|202 | ···················· | ···················· | ···················· | ···················· | ···················· | ···················· |
| B | 8672 | hCoV-19/England/CAMB-81AAF/2020\|EPI_ISL_443087\|202 | ···················· | ···················· | ···················· | ···················· | ···················· | ···················· |
| B | 8673 | hCoV-19/England/CAMB-81D00/2020\|EPI_ISL_443089\|202 | ···················· | ···················· | ···················· | ···················· | ···················· | ···················· |
| B | 8674 | hCoV-19/England/CAMB-1ABF18/2020\|EPI_ISL_444347\|20 | ···················· | ···················· | ···················· | ···················· | ···················· | ···················· |
| B | 8675 | hCoV-19/England/CAMB-1AC21E/2020\|EPI_ISL_444384\|20 | ···················· | ···················· | ···················· | ···················· | ···················· | ···················· |
| B | 8676 | hCoV-19/England/CAMB-79943/2020\|EPI_ISL_444393\|202 | ···················· | ···················· | ···················· | ···················· | ···················· | ···················· |
| B | 8677 | hCoV-19/England/CAMB-7AD19/2020\|EPI_ISL_444394\|202 | ···················· | ···················· | ···················· | ···················· | ···················· | ···················· |
| B | 8678 | hCoV-19/England/CAMB-7B219/2020\|EPI_ISL_444395\|202 | ···················· | ···················· | ···················· | ···················· | ···················· | ···················· |
| B | 8679 | hCoV-19/England/CAMB-7FBED/2020\|EPI_ISL_444397\|202 | ···················· | ···················· | ···················· | ···················· | ···················· | ···················· |
| B | 8680 | hCoV-19/England/CAMB-7FC71/2020\|EPI_ISL_444398\|202 | ···················· | ···················· | ···················· | ···················· | ···················· | ···················· |
| B | 8681 | hCoV-19/Wales/PHWC-28B3E/2020\|EPI_ISL_445405\|2020- | ···················· | ···················· | ···················· | ···················· | ···················· | ···················· |
| B | 8682 | hCoV-19/Wales/PHWC-28B4D/2020\|EPI_ISL_445406\|2020- | ···················· | ···················· | ···················· | ···················· | ···················· | ···················· |
| B | 8683 | hCoV-19/Wales/PHWC-28B6B/2020\|EPI_ISL_445408\|2020- | ···················· | ···················· | ···················· | ···················· | ···················· | ···················· |
| B | 8684 | hCoV-19/Wales/PHWC-28B7A/2020\|EPI_ISL_445409\|2020- | ···················· | ···················· | ···················· | ···················· | ···················· | ···················· |
| B | 8685 | hCoV-19/Wales/PHWC-28BA7/2020\|EPI_ISL_445412\|2020- | ···················· | ···················· | ···················· | ···················· | ···················· | ···················· |
| B | 8686 | hCoV-19/Wales/PHWC-28BB6/2020\|EPI_ISL_445413\|2020- | ···················· | ···················· | ···················· | ···················· | ···················· | ···················· |
| B | 8687 | hCoV-19/Wales/PHWC-28BC5/2020\|EPI_ISL_445414\|2020- | ···················· | ···················· | ···················· | ···················· | ···················· | ···················· |
| B | 8688 | hCoV-19/Wales/PHWC-28BE3/2020\|EPI_ISL_445416\|2020- | ···················· | ···················· | ···················· | ···················· | ···················· | ···················· |
| B | 8689 | hCoV-19/Wales/PHWC-28BF2/2020\|EPI_ISL_445417\|2020- | ···················· | ···················· | ···················· | ···················· | ···················· | ···················· |
| B | 8690 | hCoV-19/Wales/PHWC-28C1D/2020\|EPI_ISL_445419\|2020- | ···················· | ···················· | ···················· | ···················· | ···················· | ···················· |

| | | | | | | | | |
|---|---|---|---|---|---|---|---|---|
| B | 8691 hCoV-19/Wales/PHWC-28C2C/2020\|EPI_ISL_445420\|2020- | .................... | .................... | .................... | .................... | .................... | .................... | |
| B | 8692 hCoV-19/Wales/PHWC-28C4A/2020\|EPI_ISL_445422\|2020- | .................... | .................... | .................... | .................... | .................... | .................... | |
| B | 8693 hCoV-19/Wales/PHWC-28C59/2020\|EPI_ISL_445423\|2020- | .................... | .................... | .................... | .................... | .................... | .................... | |
| B | 8694 hCoV-19/Wales/PHWC-2ABE1/2020\|EPI_ISL_445678\|2020- | .................... | .................... | .................... | .................... | .................... | .................... | |
| B | 8695 hCoV-19/Wales/PHWC-32EED/2020\|EPI_ISL_446706\|2020- | .................... | .................... | .................... | .................... | .................... | .................... | |
| B | 8696 hCoV-19/Wales/PHWC-32F62/2020\|EPI_ISL_446714\|2020- | .................... | .................... | .................... | .................... | .................... | .................... | |
| B | 8697 hCoV-19/Wales/PHWC-32F71/2020\|EPI_ISL_446715\|2020- | .................... | .................... | .................... | .................... | .................... | .................... | |
| B | 8698 hCoV-19/Wales/PHWC-32FAE/2020\|EPI_ISL_446718\|2020- | .................... | ....A............... | .................... | .................... | .................... | .................... | |
| B | 8699 hCoV-19/Wales/PHWC-32FF9/2020\|EPI_ISL_446723\|2020- | .................... | .................... | .................... | .................... | .................... | .................... | |
| B | 8700 hCoV-19/Wales/PHWC-33040/2020\|EPI_ISL_446727\|2020- | .................... | .................... | .................... | .................... | .................... | .................... | |
| B | 8701 hCoV-19/Wales/PHWC-330AA/2020\|EPI_ISL_446733\|2020- | .................... | .................... | .................... | .................... | .................... | .................... | |
| B | 8702 hCoV-19/Wales/PHWC-3312F/2020\|EPI_ISL_446741\|2020- | .................... | .................... | .................... | .................... | .................... | .................... | |
| B | 8703 hCoV-19/Wales/PHWC-33769/2020\|EPI_ISL_446744\|2020- | .................... | .................... | .................... | .................... | .................... | .................... | |
| B | 8704 hCoV-19/Wales/PHWC-337B4/2020\|EPI_ISL_446749\|2020- | .................... | .................... | .................... | .................... | .................... | .................... | |
| B | 8705 hCoV-19/Scotland/CVR826/2020\|EPI_ISL_433347\|2020-0 | .................... | .................... | .................... | .................... | .................... | .................... | |
| B | 8706 hCoV-19/Scotland/CVR833/2020\|EPI_ISL_433348\|2020-0 | .................... | .................... | .................... | .................... | .................... | .................... | |
| B | 8707 hCoV-19/Scotland/CVR849/2020\|EPI_ISL_433350\|2020-0 | .................... | ....A............... | .................... | .................... | .................... | .................... | |
| B | 8708 hCoV-19/Scotland/CVR851/2020\|EPI_ISL_433351\|2020-0 | .................... | .................... | .................... | .................... | .................... | .................... | |
| B | 8709 hCoV-19/Scotland/CVR855/2020\|EPI_ISL_433353\|2020-0 | .................... | .................... | .................... | .................... | .................... | .................... | |
| B | 8710 hCoV-19/Scotland/CVR867/2020\|EPI_ISL_433355\|2020-0 | .................... | ...................A | .................... | .................... | .................... | .................... | 1 |
| B | 8711 hCoV-19/Scotland/CVR874/2020\|EPI_ISL_433356\|2020-0 | .................... | .................... | .................... | .................... | .................... | .................... | |
| B | 8712 hCoV-19/Scotland/CVR901/2020\|EPI_ISL_433357\|2020-0 | .................... | .A.................. | .................... | .................... | .................... | .................... | |
| B | 8713 hCoV-19/Scotland/CVR902/2020\|EPI_ISL_433358\|2020-0 | .................... | .A.................. | .................... | .................... | .................... | .................... | |
| B | 8714 hCoV-19/Scotland/CVR903/2020\|EPI_ISL_433359\|2020-0 | .................... | .A.................. | .................... | .................... | .................... | .................... | |
| B | 8715 hCoV-19/Scotland/CVR922/2020\|EPI_ISL_433361\|2020-0 | .................... | .................... | .................... | .................... | .................... | .................... | |
| B | 8716 hCoV-19/Scotland/CVR929/2020\|EPI_ISL_433362\|2020-0 | .................... | .................... | .................... | .................... | .................... | .................... | |
| B | 8717 hCoV-19/Scotland/CVR942/2020\|EPI_ISL_433364\|2020-0 | .................... | .................... | .................... | .................... | .................... | .................... | |
| B | 8718 hCoV-19/Scotland/CVR965/2020\|EPI_ISL_433366\|2020-0 | .................... | ...................A | .................... | .................... | .................... | .................... | 1 |
| B | 8719 hCoV-19/Scotland/CVR968/2020\|EPI_ISL_433367\|2020-0 | .................... | .................... | .................... | .................... | .................... | .................... | |
| B | 8720 hCoV-19/Scotland/CVR969/2020\|EPI_ISL_433368\|2020-0 | .................... | .................... | .................... | .................... | .................... | .................... | |
| B | 8721 hCoV-19/DRC/1377/2020\|EPI_ISL_434678\|2020-04-07 | .................... | .................... | .................... | .................... | .................... | .................... | |
| B | 8722 hCoV-19/USA/MD-HP00016/2020\|EPI_ISL_434682\|2020-03 | .................... | .................... | .................... | .................... | .................... | .................... | |
| B | 8723 hCoV-19/USA/MD-HP00020/2020\|EPI_ISL_434683\|2020-03 | .................... | .................... | .................... | .................... | .................... | .................... | |
| B | 8724 hCoV-19/USA/MD-HP00023/2020\|EPI_ISL_434684\|2020-03 | .................... | .................... | .................... | .................... | .................... | .................... | |
| B | 8725 hCoV-19/USA/MD-HP00024/2020\|EPI_ISL_434685\|2020-03 | .................... | .................... | .................... | .................... | .................... | .................... | |
| B | 8726 hCoV-19/USA/MD-HP00030/2020\|EPI_ISL_434686\|2020-03 | .................... | .................... | .................... | .................... | .................... | .................... | |
| B | 8727 hCoV-19/USA/DC-HP00006/2020\|EPI_ISL_434687\|2020-03 | .................... | .................... | .................... | .................... | .................... | .................... | |
| B | 8728 hCoV-19/USA/DC-HP00007/2020\|EPI_ISL_434688\|2020-03 | .................... | .................... | .................... | .................... | .................... | .................... | |
| B | 8729 hCoV-19/USA/MD-HP00010/2020\|EPI_ISL_434689\|2020-03 | .................... | .................... | .................... | .................... | .................... | .................... | |
| B | 8730 hCoV-19/USA/DC-HP00022/2020\|EPI_ISL_434691\|2020-03 | .................... | .................... | .................... | .................... | .................... | .................... | |
| B | 8731 hCoV-19/Thailand/NIH-59/2020\|EPI_ISL_434693\|2020-0 | .................... | .................... | .................... | .................... | .................... | .................... | |
| B | 8732 hCoV-19/Thailand/NIH-162/2020\|EPI_ISL_434694\|2020- | .................... | .................... | .................... | .................... | .................... | .................... | |
| B | 8733 hCoV-19/Thailand/NIH-1889/2020\|EPI_ISL_434697\|2020 | .................... | .................... | .................... | .................... | .................... | .................... | |
| B | 8734 hCoV-19/Thailand/NIH-2296/2020\|EPI_ISL_434699\|2020 | .................... | .................... | .................... | .................... | .................... | .................... | |
| B | 8735 hCoV-19/USA/WA-S647/2020\|EPI_ISL_438200\|2020-04-19 | .................... | .................... | .................... | .................... | .................... | .................... | |
| B | 8736 hCoV-19/USA/WA-S648/2020\|EPI_ISL_438201\|2020-04-19 | .................... | .................... | .................... | .................... | .................... | .................... | |
| B | 8737 hCoV-19/USA/WA-S649/2020\|EPI_ISL_438202\|2020-04-19 | .................... | .................... | .................... | .................... | .................... | .................... | |
| B | 8738 hCoV-19/USA/WA-S650/2020\|EPI_ISL_438203\|2020-04-20 | .................... | .................... | .................... | .................... | .................... | .................... | |
| B | 8739 hCoV-19/USA/WA-S651/2020\|EPI_ISL_438204\|2020-04-20 | .................... | .................... | .................... | .................... | .................... | .................... | |
| B | 8740 hCoV-19/USA/WA-S652/2020\|EPI_ISL_438205\|2020-04-20 | .................... | .................... | .................... | .................... | .................... | .................... | |
| B | 8741 hCoV-19/USA/WA-S653/2020\|EPI_ISL_438206\|2020-04-15 | .................... | .................... | .................... | .................... | .................... | .................... | |
| B | 8742 hCoV-19/USA/WA-S654/2020\|EPI_ISL_438207\|2020-04-21 | .................... | .................... | .................... | .................... | .................... | .................... | |
| B | 8743 hCoV-19/USA/WA-S655/2020\|EPI_ISL_438208\|2020-04-16 | .................... | .................... | .................... | .................... | .................... | .................... | |
| B | 8744 hCoV-19/USA/WA-S656/2020\|EPI_ISL_438209\|2020-04-21 | .................... | .................... | .................... | .................... | .................... | .................... | |
| B | 8745 hCoV-19/USA/WA-S657/2020\|EPI_ISL_438210\|2020-04-21 | .................... | .................... | .................... | .................... | .................... | .................... | |
| B | 8746 hCoV-19/USA/WA-S658/2020\|EPI_ISL_438211\|2020-04-21 | .................... | .................... | .................... | .................... | .................... | .................... | |
| B | 8747 hCoV-19/USA/WA-S659/2020\|EPI_ISL_438212\|2020-04-21 | .................... | .................... | .................... | .................... | .................... | .................... | |
| B | 8748 hCoV-19/USA/WA-S660/2020\|EPI_ISL_438213\|2020-04-21 | .................... | .................... | .................... | .................... | .................... | .................... | |
| B | 8749 hCoV-19/USA/WA-S661/2020\|EPI_ISL_438214\|2020-04-21 | .................... | .................... | .................... | .................... | .................... | .................... | |
| B | 8750 hCoV-19/USA/WA-S662/2020\|EPI_ISL_438215\|2020-04-20 | .................... | .................... | .................... | .................... | .................... | .................... | |
| B | 8751 hCoV-19/USA/WA-S663/2020\|EPI_ISL_438216\|2020-04-21 | .................... | .................... | .................... | .................... | .................... | .................... | |
| B | 8752 hCoV-19/USA/WA-S664/2020\|EPI_ISL_438217\|2020-04-22 | .................... | .................... | .................... | .................... | .................... | .................... | |
| B | 8753 hCoV-19/USA/WA-S665/2020\|EPI_ISL_438218\|2020-04-22 | .................... | .................... | .................... | .................... | .................... | .................... | |
| B | 8754 hCoV-19/USA/WA-S666/2020\|EPI_ISL_438219\|2020-04-22 | .................... | .................... | .................... | .................... | .................... | .................... | |
| B | 8755 hCoV-19/USA/DC-HP00072/2020\|EPI_ISL_438230\|2020-03 | .................... | .................... | .................... | .................... | .................... | .................... | |
| B | 8756 hCoV-19/USA/DC-HP00073/2020\|EPI_ISL_438231\|2020-03 | .................... | .................... | .................... | .................... | .................... | .................... | |
| B | 8757 hCoV-19/England/CAMB-75A80/2020\|EPI_ISL_438260\|202 | .................... | .................... | .................... | .................... | .................... | .................... | |
| B | 8758 hCoV-19/England/CAMB-75C8A/2020\|EPI_ISL_438262\|202 | .................... | .................... | .................... | .................... | .................... | .................... | |
| B | 8759 hCoV-19/England/CAMB-76B9B/2020\|EPI_ISL_438265\|202 | .................... | .................... | .................... | .................... | .................... | .................... | |
| B | 8760 hCoV-19/England/CAMB-76278/2020\|EPI_ISL_438266\|202 | .................... | .................... | .................... | .................... | .................... | .................... | |
| B | 8761 hCoV-19/England/CAMB-76481/2020\|EPI_ISL_438267\|202 | .................... | .................... | .................... | .................... | .................... | .................... | |
| B | 8762 hCoV-19/England/CAMB-760F6/2020\|EPI_ISL_438268\|202 | .................... | .................... | .................... | .................... | .................... | .................... | |
| B | 8763 hCoV-19/England/CAMB-76849/2020\|EPI_ISL_438269\|202 | .................... | .................... | .................... | .................... | .................... | .................... | |
| B | 8764 hCoV-19/England/CAMB-75DD2/2020\|EPI_ISL_438270\|202 | .................... | .................... | .................... | .................... | .................... | .................... | |
| B | 8765 hCoV-19/England/CAMB-7682B/2020\|EPI_ISL_438272\|202 | .................... | .................... | .................... | .................... | .................... | .................... | |
| B | 8766 hCoV-19/England/CAMB-76445/2020\|EPI_ISL_438273\|202 | .................... | .................... | .................... | .................... | .................... | .................... | |
| B | 8767 hCoV-19/England/CAMB-76393/2020\|EPI_ISL_438274\|202 | .................... | .................... | .................... | .................... | .................... | .................... | |
| B | 8768 hCoV-19/England/CAMB-75E66/2020\|EPI_ISL_438276\|202 | .................... | .................... | .................... | .................... | .................... | .................... | |

| | | | | | | | | |
|---|---|---|---|---|---|---|---|---|
| B | 8769 | hCoV-19/England/CAMB-75F90/2020\|EPI_ISL_438277\|202 | ···················· | ···················· | ···················· | ···················· | ···················· | ···················· |
| B | 8770 | hCoV-19/England/CAMB-75B7E/2020\|EPI_ISL_438278\|202 | ···················· | ···················· | ···················· | ···················· | ···················· | ···················· |
| B | 8771 | hCoV-19/England/CAMB-75992/2020\|EPI_ISL_438279\|202 | ···················· | ···················· | ···················· | ···················· | ···················· | ···················· |
| B | 8772 | hCoV-19/England/CAMB-76111/2020\|EPI_ISL_438280\|202 | ···················· | ···················· | ···················· | ···················· | ···················· | ···················· |
| B | 8773 | hCoV-19/England/CAMB-759B0/2020\|EPI_ISL_438282\|202 | ···················· | ···················· | ···················· | ···················· | ···················· | ···················· |
| B | 8774 | hCoV-19/England/CAMB-75D69/2020\|EPI_ISL_438284\|202 | ···················· | ···················· | ···················· | ···················· | ···················· | ···················· |
| B | 8775 | hCoV-19/England/CAMB-76894/2020\|EPI_ISL_438286\|202 | ···················· | ···················· | ···················· | ···················· | ···················· | ···················· |
| B | 8776 | hCoV-19/England/CAMB-76454/2020\|EPI_ISL_438287\|202 | ···················· | ···················· | ···················· | ···················· | ···················· | ···················· |
| B | 8777 | hCoV-19/England/CAMB-764EB/2020\|EPI_ISL_438288\|202 | ···················· | ···················· | ···················· | ···················· | ···················· | ···················· |
| B | 8778 | hCoV-19/England/CAMB-76375/2020\|EPI_ISL_438289\|202 | ···················· | ···················· | ···················· | ···················· | ···················· | ···················· |
| B | 8779 | hCoV-19/England/CAMB-75F63/2020\|EPI_ISL_438290\|202 | ···················· | ···················· | ···················· | ···················· | ···················· | ···················· |
| B | 8780 | hCoV-19/England/CAMB-75929/2020\|EPI_ISL_438291\|202 | ···················· | ···················· | ···················· | ···················· | ···················· | ···················· |
| B | 8781 | hCoV-19/England/CAMB-75CE4/2020\|EPI_ISL_438292\|202 | ···················· | ···················· | ···················· | ···················· | ···················· | ···················· |
| B | 8782 | hCoV-19/England/CAMB-7630C/2020\|EPI_ISL_438293\|202 | ···················· | ···················· | ···················· | ···················· | ···················· | ···················· |
| B | 8783 | hCoV-19/England/CAMB-76B8C/2020\|EPI_ISL_438294\|202 | ···················· | ···················· | ···················· | ···················· | ···················· | ···················· |
| B | 8784 | hCoV-19/England/CAMB-769CE/2020\|EPI_ISL_438295\|202 | ···················· | ···················· | ···················· | ···················· | ···················· | ···················· |
| B | 8785 | hCoV-19/England/CAMB-76B22/2020\|EPI_ISL_438296\|202 | ···················· | ···················· | ···················· | ···················· | ···················· | ···················· |
| B | 8786 | hCoV-19/England/CAMB-75B32/2020\|EPI_ISL_438297\|202 | ···················· | ···················· | ···················· | ···················· | ···················· | ···················· |
| B | 8787 | hCoV-19/England/CAMB-75BC9/2020\|EPI_ISL_438299\|202 | ···················· | ···················· | ···················· | ···················· | ···················· | ···················· |
| B | 8788 | hCoV-19/England/CAMB-776B7/2020\|EPI_ISL_439501\|202 | ···················· | ···················· | ···················· | ···················· | ···················· | ···················· |
| B | 8789 | hCoV-19/England/CAMB-779DC/2020\|EPI_ISL_439502\|202 | ···················· | ···················· | ···················· | ···················· | ···················· | ···················· |
| B | 8790 | hCoV-19/England/CAMB-774BD/2020\|EPI_ISL_439503\|202 | ···················· | ···················· | ···················· | ···················· | ···················· | ···················· |
| B | 8791 | hCoV-19/England/CAMB-76EED/2020\|EPI_ISL_439504\|202 | ···················· | ···················· | ···················· | ···················· | ···················· | ···················· |
| B | 8792 | hCoV-19/England/CAMB-76120/2020\|EPI_ISL_439505\|202 | ···················· | ···················· | ···················· | ···················· | ···················· | ···················· |
| B | 8793 | hCoV-19/England/CAMB-76032/2020\|EPI_ISL_439507\|202 | ···················· | ···················· | ···················· | ···················· | ···················· | ···················· |
| B | 8794 | hCoV-19/England/CAMB-76339/2020\|EPI_ISL_439508\|202 | ···················· | ···················· | ···················· | ···················· | ···················· | ···················· |
| B | 8795 | hCoV-19/England/CAMB-77893/2020\|EPI_ISL_439509\|202 | ···················· | ···················· | ···················· | ···················· | ···················· | ···················· |
| B | 8796 | hCoV-19/England/CAMB-77B30/2020\|EPI_ISL_439510\|202 | ···················· | ···················· | ···················· | ···················· | ···················· | ···················· |
| B | 8797 | hCoV-19/England/CAMB-76EA1/2020\|EPI_ISL_439511\|202 | ···················· | ···················· | ···················· | ···················· | ···················· | ···················· |
| B | 8798 | hCoV-19/England/CAMB-76D2C/2020\|EPI_ISL_439514\|202 | ···················· | ···················· | ···················· | ···················· | ···················· | ···················· |
| B | 8799 | hCoV-19/England/CAMB-771F2/2020\|EPI_ISL_439516\|202 | ···················· | ···················· | ···················· | ···················· | ···················· | ···················· |
| B | 8800 | hCoV-19/England/CAMB-770D7/2020\|EPI_ISL_439517\|202 | ···················· | ···················· | ···················· | ···················· | ···················· | ···················· |
| B | 8801 | hCoV-19/England/CAMB-760BA/2020\|EPI_ISL_439518\|202 | ···················· | ···················· | ···················· | ···················· | ···················· | ···················· |
| B | 8802 | hCoV-19/England/CAMB-77A15/2020\|EPI_ISL_439519\|202 | ···················· | ···················· | ···················· | ···················· | ···················· | ···················· |
| B | 8803 | hCoV-19/England/CAMB-765CA/2020\|EPI_ISL_439520\|202 | ···················· | ···················· | ···················· | ···················· | ···················· | ···················· |
| B | 8804 | hCoV-19/England/CAMB-76ECF/2020\|EPI_ISL_439521\|202 | ···················· | ···················· | ···················· | ···················· | ···················· | ···················· |
| B | 8805 | hCoV-19/England/CAMB-766B8/2020\|EPI_ISL_439522\|202 | ···················· | ···················· | ···················· | ···················· | ···················· | ···················· |
| B | 8806 | hCoV-19/England/CAMB-7607E/2020\|EPI_ISL_439523\|202 | ···················· | ···················· | ···················· | ···················· | ···················· | ···················· |
| B | 8807 | hCoV-19/England/CAMB-77990/2020\|EPI_ISL_439524\|202 | ···················· | ···················· | ···················· | ···················· | ···················· | ···················· |
| B | 8808 | hCoV-19/England/CAMB-76D68/2020\|EPI_ISL_439525\|202 | ···················· | ···················· | ···················· | ···················· | ···················· | ···················· |
| B | 8809 | hCoV-19/England/CAMB-77B5E/2020\|EPI_ISL_439526\|202 | ···················· | ···················· | ···················· | ···················· | ···················· | ···················· |
| B | 8810 | hCoV-19/England/CAMB-760C9/2020\|EPI_ISL_439527\|202 | ···················· | ···················· | ···················· | ···················· | ···················· | ···················· |
| B | 8811 | hCoV-19/England/CAMB-7781B/2020\|EPI_ISL_439528\|202 | ···················· | ···················· | ···················· | ···················· | ···················· | ···················· |
| B | 8812 | hCoV-19/England/CAMB-71D03/2020\|EPI_ISL_439532\|202 | ···················· | ···················· | ···················· | ···················· | ···················· | ···················· |
| B | 8813 | hCoV-19/England/CAMB-72616/2020\|EPI_ISL_439533\|202 | ···················· | ···················· | ···················· | ···················· | ···················· | ···················· |
| B | 8814 | hCoV-19/England/CAMB-71BDC/2020\|EPI_ISL_439534\|202 | ···················· | ···················· | ···················· | ···················· | ···················· | ···················· |
| B | 8815 | hCoV-19/England/CAMB-72379/2020\|EPI_ISL_439535\|202 | ···················· | ···················· | ···················· | ···················· | ···················· | ···················· |
| B | 8816 | hCoV-19/England/CAMB-71B81/2020\|EPI_ISL_439536\|202 | ···················· | ···················· | ···················· | ···················· | ···················· | ···················· |
| B | 8817 | hCoV-19/England/CAMB-727C8/2020\|EPI_ISL_439538\|202 | ···················· | ···················· | ···················· | ···················· | ···················· | ···················· |
| B | 8818 | hCoV-19/England/CAMB-72494/2020\|EPI_ISL_439540\|202 | ···················· | ···················· | ···················· | ···················· | ···················· | ···················· |
| B | 8819 | hCoV-19/England/CAMB-72546/2020\|EPI_ISL_439541\|202 | ···················· | ···················· | ···················· | ···················· | ···················· | ···················· |
| B | 8820 | hCoV-19/England/CAMB-7240D/2020\|EPI_ISL_439542\|202 | ···················· | ···················· | ···················· | ···················· | ···················· | ···················· |
| B | 8821 | hCoV-19/England/CAMB-7225E/2020\|EPI_ISL_439545\|202 | ···················· | ·············A······ | ···················· | ···················· | ···················· | ···················· |
| B | 8822 | hCoV-19/England/CAMB-723E2/2020\|EPI_ISL_439547\|202 | ···················· | ···················· | ···················· | ···················· | ···················· | ···················· |
| B | 8823 | hCoV-19/England/CAMB-72018/2020\|EPI_ISL_439548\|202 | ···················· | ···················· | ···················· | ···················· | ···················· | ···················· |
| B | 8824 | hCoV-19/England/CAMB-72959/2020\|EPI_ISL_439562\|202 | ···················· | ···················· | ···················· | ···················· | ···················· | ···················· |
| B | 8825 | hCoV-19/England/CAMB-71CD9/2020\|EPI_ISL_439590\|202 | ···················· | ···················· | ···················· | ···················· | ···················· | ···················· |
| B | 8826 | hCoV-19/England/CAMB-7284D/2020\|EPI_ISL_439591\|202 | ···················· | ···················· | ···················· | ···················· | ···················· | ···················· |
| B | 8827 | hCoV-19/England/CAMB-71F1C/2020\|EPI_ISL_439592\|202 | ···················· | ···················· | ···················· | ···················· | ···················· | ···················· |
| B | 8828 | hCoV-19/England/CAMB-7294A/2020\|EPI_ISL_439594\|202 | ···················· | ···················· | ···················· | ···················· | ···················· | ···················· |
| B | 8829 | hCoV-19/England/CAMB-72485/2020\|EPI_ISL_439596\|202 | ···················· | ···················· | ···················· | ···················· | ···················· | ···················· |
| B | 8830 | hCoV-19/England/CAMB-7218E/2020\|EPI_ISL_439598\|202 | ···················· | ···················· | ···················· | ···················· | ···················· | ···················· |
| B | 8831 | hCoV-19/England/CAMB-71D7C/2020\|EPI_ISL_439599\|202 | ···················· | ···················· | ···················· | ···················· | ···················· | ···················· |
| B | 8832 | hCoV-19/England/CAMB-84AE8/2020\|EPI_ISL_440501\|202 | ···················· | ···················· | ···················· | ···················· | ···················· | ···················· |
| B | 8833 | hCoV-19/England/CAMB-84BE5/2020\|EPI_ISL_440503\|202 | ···················· | ···················· | ···················· | ···················· | ···················· | ···················· |
| B | 8834 | hCoV-19/England/CAMB-84787/2020\|EPI_ISL_440506\|202 | ···················· | ···················· | ···················· | ···················· | ···················· | ···················· |
| B | 8835 | hCoV-19/England/CAMB-84839/2020\|EPI_ISL_440507\|202 | ···················· | ···················· | ···················· | ···················· | ···················· | ···················· |
| B | 8836 | hCoV-19/England/CAMB-84C2E/2020\|EPI_ISL_440510\|202 | ···················· | ···················· | ···················· | ···················· | ···················· | ···················· |
| B | 8837 | hCoV-19/England/CAMB-84C6A/2020\|EPI_ISL_440511\|202 | ···················· | ···················· | ···················· | ···················· | ···················· | ···················· |
| B | 8838 | hCoV-19/England/CAMB-84BD6/2020\|EPI_ISL_440515\|202 | ···················· | ···················· | ···················· | ···················· | ···················· | ···················· |
| B | 8839 | hCoV-19/England/CAMB-84B8B/2020\|EPI_ISL_440516\|202 | ···················· | ···················· | ···················· | ···················· | ···················· | ···················· |
| B | 8840 | hCoV-19/England/CAMB-84848/2020\|EPI_ISL_440518\|202 | ···················· | ···················· | ···················· | ···················· | ···················· | ···················· |
| B | 8841 | hCoV-19/England/CAMB-84A9D/2020\|EPI_ISL_440519\|202 | ···················· | ···················· | ···················· | ···················· | ···················· | ···················· |
| B | 8842 | hCoV-19/England/CAMB-8482A/2020\|EPI_ISL_440521\|202 | ···················· | ···················· | ···················· | ···················· | ···················· | ···················· |
| B | 8843 | hCoV-19/England/CAMB-84B6D/2020\|EPI_ISL_440525\|202 | ···················· | ···················· | ···················· | ···················· | ···················· | ···················· |
| B | 8844 | hCoV-19/England/CAMB-84BC7/2020\|EPI_ISL_440526\|202 | ···················· | ···················· | ···················· | ···················· | ···················· | ···················· |
| B | 8845 | hCoV-19/England/CAMB-84C3D/2020\|EPI_ISL_440547\|202 | ···················· | ···················· | ···················· | ···················· | ···················· | ···················· |
| B | 8846 | hCoV-19/England/CAMB-7A3CC/2020\|EPI_ISL_440574\|202 | ···················· | ···················· | ···················· | ···················· | ···················· | ···················· |

| | | | | | | | |
|---|---|---|---|---|---|---|---|
| B | 8847 hCoV-19/England/CAMB-79CE0/2020\|EPI_ISL_440576\|202 | ···················· | ···················· | ···················· | ···················· | ···················· | ···················· |
| B | 8848 hCoV-19/England/CAMB-79DA1/2020\|EPI_ISL_440577\|202 | ···················· | ···················· | ···················· | ···················· | ···················· | ···················· |
| B | 8849 hCoV-19/England/CAMB-78DEE/2020\|EPI_ISL_440579\|202 | ···················· | ···················· | ···················· | ···················· | ···················· | ···················· |
| B | 8850 hCoV-19/England/CAMB-7A1B3/2020\|EPI_ISL_440581\|202 | ···················· | ···················· | ···················· | ···················· | ···················· | ···················· |
| B | 8851 hCoV-19/England/CAMB-7A7EE/2020\|EPI_ISL_440582\|202 | ···················· | ···················· | ···················· | ···················· | ···················· | ···················· |
| B | 8852 hCoV-19/England/CAMB-7A21A/2020\|EPI_ISL_440583\|202 | ···················· | ···················· | ···················· | ···················· | ···················· | ···················· |
| B | 8853 hCoV-19/England/CAMB-7A827/2020\|EPI_ISL_440584\|202 | ···················· | ···················· | ···················· | ···················· | ···················· | ···················· |
| B | 8854 hCoV-19/England/CAMB-7A159/2020\|EPI_ISL_440586\|202 | ···················· | ···················· | ···················· | ···················· | ···················· | ···················· |
| B | 8855 hCoV-19/England/CAMB-7A11D/2020\|EPI_ISL_440589\|202 | ···················· | ···················· | ···················· | ···················· | ···················· | ···················· |
| B | 8856 hCoV-19/England/CAMB-7A4C9/2020\|EPI_ISL_440590\|202 | ···················· | ···················· | ···················· | ···················· | ···················· | ···················· |
| B | 8857 hCoV-19/England/CAMB-7A7FD/2020\|EPI_ISL_440591\|202 | ···T················ | ···················· | ···················· | ···················· | ···················· | ···················· |
| B | 8858 hCoV-19/England/CAMB-79D74/2020\|EPI_ISL_440592\|202 | ···················· | ···················· | ···················· | ···················· | ···················· | ···················· |
| B | 8859 hCoV-19/England/CAMB-7A46F/2020\|EPI_ISL_440593\|202 | ···················· | ···················· | ···················· | ···················· | ···················· | ···················· |
| B | 8860 hCoV-19/England/CAMB-7A10E/2020\|EPI_ISL_440595\|202 | ···················· | ···················· | ···················· | ···················· | ···················· | ···················· |
| B | 8861 hCoV-19/England/CAMB-79E44/2020\|EPI_ISL_440597\|202 | ···················· | ···················· | ···················· | ···················· | ···················· | ···················· |
| B | 8862 hCoV-19/England/CAMB-7B695/2020\|EPI_ISL_440598\|202 | ···················· | ···················· | ···················· | ···················· | ···················· | ···················· |
| B | 8863 hCoV-19/England/CAMB-79CC2/2020\|EPI_ISL_440599\|202 | ···················· | ···················· | ···················· | ···················· | R··················· | ···················· |
| B | 8864 hCoV-19/England/CAMB-7A57B/2020\|EPI_ISL_441800\|202 | ···················· | ···················· | ···················· | ···················· | ···················· | ···················· |
| B | 8865 hCoV-19/England/CAMB-79011/2020\|EPI_ISL_441801\|202 | ···················· | ···················· | ···················· | ···················· | ···················· | ···················· |
| B | 8866 hCoV-19/England/CAMB-78CE1/2020\|EPI_ISL_441802\|202 | ···················· | ···················· | ···················· | ···················· | ···················· | ···················· |
| B | 8867 hCoV-19/England/CAMB-7804F/2020\|EPI_ISL_441803\|202 | ···················· | ···················· | ···················· | ···················· | ···················· | ···················· |
| B | 8868 hCoV-19/England/CAMB-78D75/2020\|EPI_ISL_441804\|202 | ···················· | ···················· | ···················· | ···················· | ···················· | ···················· |
| B | 8869 hCoV-19/England/CAMB-7A6C3/2020\|EPI_ISL_441806\|202 | ···················· | ···················· | ···················· | ···················· | ···················· | ···················· |
| B | 8870 hCoV-19/England/CAMB-79882/2020\|EPI_ISL_441809\|202 | ···················· | ···················· | ···················· | ···················· | ···················· | ···················· |
| B | 8871 hCoV-19/England/CAMB-79864/2020\|EPI_ISL_441811\|202 | ···················· | ···················· | ···················· | ···················· | ···················· | ···················· |
| B | 8872 hCoV-19/England/CAMB-7905D/2020\|EPI_ISL_441813\|202 | ···················· | ···················· | ···················· | ···················· | ···················· | ···················· |
| B | 8873 hCoV-19/England/CAMB-797D0/2020\|EPI_ISL_441816\|202 | ···················· | ···················· | ···················· | ···················· | ···················· | ···················· |
| B | 8874 hCoV-19/England/CAMB-78FF7/2020\|EPI_ISL_441817\|202 | ···················· | ···················· | ···················· | ···················· | ···················· | ···················· |
| B | 8875 hCoV-19/England/CAMB-79846/2020\|EPI_ISL_441818\|202 | ···················· | ···················· | ···················· | ···················· | ···················· | ···················· |
| B | 8876 hCoV-19/England/CAMB-79512/2020\|EPI_ISL_441820\|202 | ···················· | ···················· | ···················· | ···················· | ···················· | ···················· |
| B | 8877 hCoV-19/England/CAMB-7A960/2020\|EPI_ISL_441824\|202 | ···················· | ···················· | ···················· | ···················· | ···················· | ···················· |
| B | 8878 hCoV-19/England/CAMB-791C3/2020\|EPI_ISL_441826\|202 | ···················· | ···················· | ···················· | ···················· | ···················· | ···················· |
| B | 8879 hCoV-19/England/CAMB-78D1B/2020\|EPI_ISL_441827\|202 | ···················· | ···················· | ···················· | ···················· | ···················· | ···················· |
| B | 8880 hCoV-19/England/CAMB-7A47E/2020\|EPI_ISL_441830\|202 | ···················· | ···················· | ···················· | ···················· | ···················· | ···················· |
| B | 8881 hCoV-19/England/CAMB-7813D/2020\|EPI_ISL_441831\|202 | ···················· | ···················· | ···················· | ···················· | ···················· | ···················· |
| B | 8882 hCoV-19/England/CAMB-79390/2020\|EPI_ISL_441832\|202 | ···················· | ···················· | ···················· | ···················· | ···················· | ···················· |
| B | 8883 hCoV-19/England/CAMB-793AF/2020\|EPI_ISL_441833\|202 | ···T················ | ···················· | ···················· | ···················· | ···················· | ···················· |
| B | 8884 hCoV-19/England/CAMB-780D6/2020\|EPI_ISL_441835\|202 | ···················· | ···················· | ···················· | ···················· | ···················· | ···················· |
| B | 8885 hCoV-19/England/CAMB-77E82/2020\|EPI_ISL_441836\|202 | ···················· | ···················· | ···················· | ···················· | ···················· | ···················· |
| B | 8886 hCoV-19/England/CAMB-77EBF/2020\|EPI_ISL_441837\|202 | ···················· | ···················· | ···················· | ···················· | ···················· | ···················· |
| B | 8887 hCoV-19/England/CAMB-7980A/2020\|EPI_ISL_441841\|202 | ···················· | ···················· | ···················· | ···················· | ···················· | ···················· |
| B | 8888 hCoV-19/England/CAMB-77D3A/2020\|EPI_ISL_441842\|202 | ···················· | ···················· | ···················· | ···················· | ···················· | ···················· |
| B | 8889 hCoV-19/England/CAMB-796A6/2020\|EPI_ISL_441843\|202 | ···················· | ···················· | ···················· | ···················· | ···················· | ···················· |
| B | 8890 hCoV-19/England/CAMB-7A2FC/2020\|EPI_ISL_441844\|202 | ···················· | ···················· | ···················· | ···················· | ···················· | ···················· |
| B | 8891 hCoV-19/England/CAMB-7A5F3/2020\|EPI_ISL_441845\|202 | ···················· | ···················· | ···················· | ···················· | ···················· | ···················· |
| B | 8892 hCoV-19/England/NOTT-1108AA/2020\|EPI_ISL_441872\|20 | ···················· | ···················· | ···················· | ···················· | ···················· | ···················· |
| B | 8893 hCoV-19/England/NOTT-1108B9/2020\|EPI_ISL_441873\|20 | ···················· | ···················· | ···················· | ···················· | ···················· | ···················· |
| B | 8894 hCoV-19/England/NOTT-110989/2020\|EPI_ISL_441878\|20 | ···················· | ···················· | ···················· | ···················· | ···················· | ···················· |
| B | 8895 hCoV-19/England/NOTT-110998/2020\|EPI_ISL_441879\|20 | ···················· | ···················· | ···················· | ···················· | ···················· | ···················· |
| B | 8896 hCoV-19/England/BRIS-124FAE/2020\|EPI_ISL_444000\|20 | ···················· | ···················· | ···················· | ···················· | ···················· | ···················· |
| B | 8897 hCoV-19/England/BRIS-1253A1/2020\|EPI_ISL_444001\|20 | ···················· | ···················· | ···················· | ···················· | ···················· | ···················· |
| B | 8898 hCoV-19/England/BRIS-125013/2020\|EPI_ISL_444002\|20 | ···················· | ···················· | ···················· | ···················· | ···················· | ···················· |
| B | 8899 hCoV-19/England/BRIS-1251D4/2020\|EPI_ISL_444003\|20 | ···················· | ···················· | ···················· | ···················· | ···················· | ···················· |
| B | 8900 hCoV-19/England/BRIS-124867/2020\|EPI_ISL_444009\|20 | ···················· | ···················· | ···················· | ···················· | ···················· | ···················· |
| B | 8901 hCoV-19/England/BRIS-12480D/2020\|EPI_ISL_444010\|20 | ···················· | ···················· | ···················· | ···················· | ···················· | ···················· |
| B | 8902 hCoV-19/England/BRIS-1256B7/2020\|EPI_ISL_444011\|20 | ···················· | ···················· | ···················· | ···················· | ···················· | ···················· |
| B | 8903 hCoV-19/England/BRIS-125268/2020\|EPI_ISL_444012\|20 | ···················· | ···················· | ···················· | ···················· | ···················· | ···················· |
| B | 8904 hCoV-19/England/BRIS-125AD9/2020\|EPI_ISL_444013\|20 | ···················· | ···················· | ···················· | ···················· | ···················· | ···················· |
| B | 8905 hCoV-19/England/BRIS-124B5F/2020\|EPI_ISL_444015\|20 | ···················· | ···················· | ···················· | ···················· | ···················· | ···················· |
| B | 8906 hCoV-19/England/BRIS-124D2C/2020\|EPI_ISL_444016\|20 | ···················· | ···················· | ···················· | ···················· | ···················· | ···················· |
| B | 8907 hCoV-19/USA/CA-CZB-1105/2020\|EPI_ISL_444063\|2020-0 | ···················· | ···················· | ···················· | ···················· | ···················· | ···················· |
| B | 8908 hCoV-19/USA/CA-CZB-1107/2020\|EPI_ISL_444065\|2020-0 | ···················· | ···················· | ···················· | ···················· | ···················· | ···················· |
| B | 8909 hCoV-19/USA/CA-CZB-1108/2020\|EPI_ISL_444066\|2020-0 | ···················· | ···················· | ···················· | ···················· | ···················· | ···················· |
| B | 8910 hCoV-19/USA/CA-CZB-1109/2020\|EPI_ISL_444067\|2020-0 | ···················· | ···················· | ···················· | ···················· | ···················· | ···················· |
| B | 8911 hCoV-19/USA/CA-CZB-1110/2020\|EPI_ISL_444068\|2020-0 | ···················· | ···················· | ···················· | ···················· | ···················· | ···················· |
| B | 8912 hCoV-19/USA/CA-CZB-1111/2020\|EPI_ISL_444069\|2020-0 | ···················· | ···················· | ···················· | ···················· | ···················· | ···················· |
| B | 8913 hCoV-19/USA/CA-CZB-1112/2020\|EPI_ISL_444070\|2020-0 | ···················· | ···················· | ···················· | ···················· | ···················· | ···················· |
| B | 8914 hCoV-19/USA/CA-CZB-1113/2020\|EPI_ISL_444071\|2020-0 | ···················· | ···················· | ···················· | ···················· | ···················· | ···················· |
| B | 8915 hCoV-19/USA/CA-CZB-1115/2020\|EPI_ISL_444073\|2020-0 | ···················· | ···················· | ···················· | ···················· | ···················· | ···················· |
| B | 8916 hCoV-19/USA/CA-CZB-1116/2020\|EPI_ISL_444074\|2020-0 | ···················· | ···················· | ···················· | ···················· | ···················· | ···················· |
| B | 8917 hCoV-19/USA/CA-CZB-1117/2020\|EPI_ISL_444075\|2020-0 | ···················· | ···················· | ···················· | ···················· | ···················· | ···················· |
| B | 8918 hCoV-19/USA/CA-CZB-1118/2020\|EPI_ISL_444076\|2020-0 | ···················· | ···················· | ···················· | ···················· | ···················· | ···················· |
| B | 8919 hCoV-19/USA/CA-CZB-1119/2020\|EPI_ISL_444077\|2020-0 | ···················· | ···················· | ···················· | ···················· | ···················· | ···················· |
| B | 8920 hCoV-19/USA/CA-CZB-1120/2020\|EPI_ISL_444078\|2020-0 | ···················· | ···················· | ···················· | ···················· | ···················· | ···················· |
| B | 8921 hCoV-19/England/LOND-D3FCD/2020\|EPI_ISL_444079\|202 | ···················· | ···················· | ···················· | ···················· | ···················· | ···················· |
| B | 8922 hCoV-19/England/LOND-D3FDC/2020\|EPI_ISL_444080\|202 | ···················· | ···················· | ···················· | ···················· | ···················· | ···················· |
| B | 8923 hCoV-19/England/LOND-D3FEB/2020\|EPI_ISL_444081\|202 | ···················· | ···················· | ···················· | ···················· | ···················· | ···················· |
| B | 8924 hCoV-19/England/LOND-D3FFA/2020\|EPI_ISL_444082\|202 | ···················· | ···················· | ···················· | ···················· | ···················· | ···················· |

```
B 8925 hCoV-19/England/LOND-D4005/2020|EPI_ISL_444083|202 .................... .................... .................... .................... .................... ....................
B 8926 hCoV-19/England/LOND-D4014/2020|EPI_ISL_444084|202 .................... .................... .................... .................... .................... ....................
B 8927 hCoV-19/England/LOND-D4023/2020|EPI_ISL_444085|202 .................... .................... .................... .................... .................... ....................
B 8928 hCoV-19/England/LOND-D4032/2020|EPI_ISL_444086|202 .................... .................... .................... .................... .................... ....................
B 8929 hCoV-19/England/LOND-D4041/2020|EPI_ISL_444087|202 .................... .................... .................... .................... .................... ....................
B 8930 hCoV-19/England/LOND-D4050/2020|EPI_ISL_444088|202 .................... .................... .................... .................... .................... ....................
B 8931 hCoV-19/England/LOND-D406F/2020|EPI_ISL_444089|202 .................... .................... .................... .................... .................... ....................
B 8932 hCoV-19/England/LOND-D407E/2020|EPI_ISL_444090|202 .................... .................... .................... .................... .................... ....................
B 8933 hCoV-19/England/LOND-D408D/2020|EPI_ISL_444091|202 .................... .................... .................... .................... .................... ....................
B 8934 hCoV-19/England/LOND-D416C/2020|EPI_ISL_444092|202 .................... .................... .................... .................... .................... ....................
B 8935 hCoV-19/England/LOND-D417B/2020|EPI_ISL_444093|202 .................... .................... .................... .................... .................... ....................
B 8936 hCoV-19/England/LOND-D418A/2020|EPI_ISL_444094|202 .................... .................... .................... .................... .................... ....................
B 8937 hCoV-19/England/LOND-D4199/2020|EPI_ISL_444095|202 .................... .................... .................... .................... .................... ....................
B 8938 hCoV-19/England/LOND-D41A8/2020|EPI_ISL_444096|202 .................... .................... .................... .................... .................... ....................
B 8939 hCoV-19/England/LOND-D41B7/2020|EPI_ISL_444097|202 .................... .................... .................... .................... .................... ....................
B 8940 hCoV-19/England/LOND-D41C6/2020|EPI_ISL_444098|202 .................... .................... .................... .................... .................... ....................
B 8941 hCoV-19/England/LOND-D41D5/2020|EPI_ISL_444099|202 .................... .................... .................... .................... .................... ....................
B 8942 hCoV-19/Chile/RM-ISPCH-47/2020|EPI_ISL_445326|2020 .................... .................... .................... .................... .................... ....................
B 8943 hCoV-19/Chile/RM-ISPCH-48/2020|EPI_ISL_445327|2020 .................... .................... .................... .................... .................... ....................
B 8944 hCoV-19/Chile/RM-ISPCH-49/2020|EPI_ISL_445328|2020 .................... .................... .................... .................... .................... ....................
B 8945 hCoV-19/Chile/RM-ISPCH-50/2020|EPI_ISL_445329|2020 .................... .................... .................... .................... .................... ....................
B 8946 hCoV-19/Chile/RM-ISPCH-51/2020|EPI_ISL_445330|2020 .................... .................... .................... .................... .................... ....................
B 8947 hCoV-19/Chile/BI-ISPCH-2/2020|EPI_ISL_445331|2020- .................... .................... .................... .................... .................... ....................
B 8948 hCoV-19/Chile/BI-ISPCH-3/2020|EPI_ISL_445332|2020- .................... .................... .................... .................... .................... ....................
B 8949 hCoV-19/Chile/BI-ISPCH-2-2/2020|EPI_ISL_445333|202 .................... .................... .................... .................... .................... ....................
B 8950 hCoV-19/Chile/LL-ISPCH-2/2020|EPI_ISL_445334|2020- .................... .................... .................... .................... .................... ....................
B 8951 hCoV-19/Chile/LL-ISPCH-3/2020|EPI_ISL_445335|2020- .................... .................... .................... .................... .................... ....................
B 8952 hCoV-19/Chile/AR-ISPCH-5/2020|EPI_ISL_445340|2020- .................... ....A............... .................... .................... .................... ....................
B 8953 hCoV-19/Chile/AR-ISPCH-7/2020|EPI_ISL_445342|2020- .................... .................... .................... .................... .................... ....................
B 8954 hCoV-19/Chile/AR-ISPCH-8/2020|EPI_ISL_445343|2020- .................... .................... .................... .................... .................... ....................
B 8955 hCoV-19/Chile/AR-ISPCH-10/2020|EPI_ISL_445344|2020 .................... .................... .................... .................... .................... ....................
B 8956 hCoV-19/Chile/AR-ISPCH-11/2020|EPI_ISL_445345|2020 .................... .................... .................... .................... .................... ....................
B 8957 hCoV-19/Chile/AR-ISPCH-12/2020|EPI_ISL_445346|2020 .................... .................... .................... .................... .................... ....................
B 8958 hCoV-19/Chile/AR-ISPCH-13/2020|EPI_ISL_445347|2020 .................... .................... .................... .................... .................... ....................
B 8959 hCoV-19/Wales/PHWC-28A6E/2020|EPI_ISL_445392|2020- .................... .................... .................... .................... .................... ....................
B 8960 hCoV-19/Wales/PHWC-28AC8/2020|EPI_ISL_445398|2020- .................... .................... .................... .................... .................... ....................
B 8961 hCoV-19/Wales/PHWC-273CE/2020|EPI_ISL_432214|2020- .................... .................... .................... .................... .................... ....................
B 8962 hCoV-19/Wales/PHWC-29DCE/2020|EPI_ISL_432215|2020- .................... .................... .................... .................... .................... ....................
B 8963 hCoV-19/Wales/PHWC-2AF03/2020|EPI_ISL_432216|2020- .................... .................... .................... .................... .................... ....................
B 8964 hCoV-19/Wales/PHWC-27A32/2020|EPI_ISL_432217|2020- .................... .................... .................... .................... .................... ....................
B 8965 hCoV-19/Wales/PHWC-29BB5/2020|EPI_ISL_432218|2020- .................... .................... .................... .................... .................... ....................
B 8966 hCoV-19/Wales/PHWC-29C0D/2020|EPI_ISL_432219|2020- .................... .................... .................... .................... .................... ....................
B 8967 hCoV-19/Wales/PHWC-25375/2020|EPI_ISL_432220|2020- .................... .................... .................... .................... .................... ....................
B 8968 hCoV-19/Wales/PHWC-27917/2020|EPI_ISL_432221|2020- .................... .................... .................... .................... .................... ....................
B 8969 hCoV-19/Wales/PHWC-27B3F/2020|EPI_ISL_432222|2020- .................... .................... .................... .................... .................... ....................
B 8970 hCoV-19/Wales/PHWC-27443/2020|EPI_ISL_432223|2020- .................... .................... .................... .................... .................... ....................
B 8971 hCoV-19/Wales/PHWC-27786/2020|EPI_ISL_432224|2020- .................... .................... .................... .................... .................... ....................
B 8972 hCoV-19/Wales/PHWC-2B254/2020|EPI_ISL_432225|2020- .................... ...A................ .................... .................... .................... ....................
B 8973 hCoV-19/Wales/PHWC-29DEC/2020|EPI_ISL_432226|2020- .................... .................... .................... .................... .................... ....................
B 8974 hCoV-19/Wales/PHWC-27AAB/2020|EPI_ISL_432227|2020- .................... .................... .................... .................... .................... ....................
B 8975 hCoV-19/Wales/PHWC-2B281/2020|EPI_ISL_432228|2020- .................... .................... .................... .................... .................... ....................
B 8976 hCoV-19/Wales/PHWC-29D91/2020|EPI_ISL_432229|2020- .................... .................... .................... .................... .................... ....................
B 8977 hCoV-19/Wales/PHWC-27391/2020|EPI_ISL_432232|2020- .................... .................... .................... .................... .................... ....................
B 8978 hCoV-19/Scotland/CVR694/2020|EPI_ISL_433326|2020-0 .................... .................... .................... .................... .................... ....................
B 8979 hCoV-19/Scotland/CVR702/2020|EPI_ISL_433327|2020-0 .................... .................... .................... .................... .................... ....................
B 8980 hCoV-19/Scotland/CVR705/2020|EPI_ISL_433329|2020-0 .................... .................... .................... .................... .................... ....................
B 8981 hCoV-19/Scotland/CVR712/2020|EPI_ISL_433330|2020-0 .................... .A.................. .................... .................... .................... ....................
B 8982 hCoV-19/Scotland/CVR749/2020|EPI_ISL_433333|2020-0 .................... ...................A .................... .................... .................... .................... 1
B 8983 hCoV-19/Scotland/CVR752/2020|EPI_ISL_433334|2020-0 .................... .................... .................... .................... .................... ....................
B 8984 hCoV-19/Scotland/CVR754/2020|EPI_ISL_433335|2020-0 .................... .................... .................... .................... .................... ....................
B 8985 hCoV-19/Scotland/CVR780/2020|EPI_ISL_433338|2020-0 .................... .A.................. .................... .................... .................... ....................
B 8986 hCoV-19/Scotland/CVR785/2020|EPI_ISL_433339|2020-0 .................... .A.................. .................... .................... .................... ....................
B 8987 hCoV-19/Scotland/CVR788/2020|EPI_ISL_433340|2020-0 .................... .................... .................... .................... .................... ....................
B 8988 hCoV-19/Scotland/CVR796/2020|EPI_ISL_433341|2020-0 .................... .A.................. .................... .................... .................... ....................
B 8989 hCoV-19/Scotland/CVR800/2020|EPI_ISL_433342|2020-0 .................... .................... .................... .................... .................... ....................
B 8990 hCoV-19/Scotland/CVR803/2020|EPI_ISL_433344|2020-0 .................... .................... .................... .................... .................... ....................
B 8991 hCoV-19/Scotland/CVR811/2020|EPI_ISL_433346|2020-0 .................... .A.................. .................... .................... .................... ....................
B 8992 hCoV-19/Scotland/CVR919/2020|EPI_ISL_433360|2020-0 .................... .A.................. .................... .................... .................... ....................
B 8993 hCoV-19/Scotland/CVR977/2020|EPI_ISL_433370|2020-0 .................... .................... .................... .................... .................... ....................
B 8994 hCoV-19/Scotland/CVR987/2020|EPI_ISL_433371|2020-0 .................... .A.................. .................... .................... .................... ....................
B 8995 hCoV-19/Scotland/CVR997/2020|EPI_ISL_433372|2020-0 .................... .................... .................... .................... .................... ....................
B 8996 hCoV-19/Scotland/EDB1001/2020|EPI_ISL_433374|2020- .................... .................... .................... .................... .................... ....................
B 8997 hCoV-19/Scotland/EDB1005/2020|EPI_ISL_433375|2020- .................... .................... .................... .................... .................... ....................
B 8998 hCoV-19/Scotland/EDB1037/2020|EPI_ISL_433376|2020- .................... .................... .................... .................... .................... ....................
B 8999 hCoV-19/Scotland/EDB1040/2020|EPI_ISL_433377|2020- .................... .A.................. .................... .................... .................... ....................
B 9000 hCoV-19/Scotland/EDB1046/2020|EPI_ISL_433378|2020- .................... .................... .................... .................... .................... ....................
B 9001 hCoV-19/Scotland/EDB1062/2020|EPI_ISL_433379|2020- .................... .................... .................... .................... .................... ....................
B 9002 hCoV-19/Scotland/EDB1070/2020|EPI_ISL_433380|2020- .................... .A.................. .................... .................... .................... ....................
```

```
B 9003 hCoV-19/Scotland/EDB1071/2020|EPI_ISL_433381|2020- ···················· ···················· ···················· ···················· ···················· ····················
B 9004 hCoV-19/Scotland/EDB1072/2020|EPI_ISL_433382|2020- ···················· ···················· ···················· ···················· ···················· NN··················
B 9005 hCoV-19/Scotland/EDB1073/2020|EPI_ISL_433383|2020- ···················· ···················· ···················· ···················· ···················· ····················
B 9006 hCoV-19/Scotland/EDB1076/2020|EPI_ISL_433384|2020- ···················· ···················· ···················· ···················· ···················· ····················
B 9007 hCoV-19/Scotland/EDB1077/2020|EPI_ISL_433385|2020- ···················· ···················· ···················· ···················· ···················· ····················
B 9008 hCoV-19/Scotland/EDB1078/2020|EPI_ISL_433386|2020- ···················· ···················· ···················· ···················· ···················· ····················
B 9009 hCoV-19/Scotland/EDB1079/2020|EPI_ISL_433387|2020- ···················· ···················· ···················· ···················· ···················· ····················
B 9010 hCoV-19/Scotland/EDB1081/2020|EPI_ISL_433388|2020- ···················· ·A·················· ···················· ···················· ···················· ····················
B 9011 hCoV-19/Scotland/EDB1093/2020|EPI_ISL_433389|2020- ···················· ···················· ···················· ···················· ···················· ····················
B 9012 hCoV-19/Scotland/EDB1094/2020|EPI_ISL_433390|2020- ···················· ·A·················· ···················· ···················· ···················· ····················
B 9013 hCoV-19/Scotland/EDB1095/2020|EPI_ISL_433391|2020- ···················· ···················· ···················· ···················· ···················· ····················
B 9014 hCoV-19/Scotland/EDB1096/2020|EPI_ISL_433392|2020- ···················· ···················· ···················· ···················· ···················· ····················
B 9015 hCoV-19/Scotland/EDB1098/2020|EPI_ISL_433393|2020- ···················· ···················· ···················· ···················· ···················· ····················
B 9016 hCoV-19/Scotland/EDB1101/2020|EPI_ISL_433394|2020- ···················· ·A·················· ···················· ···················· ···················· ····················
B 9017 hCoV-19/Scotland/EDB1102/2020|EPI_ISL_433395|2020- ···················· ···················· ···················· ···················· ···················· ····················
B 9018 hCoV-19/Scotland/EDB1105/2020|EPI_ISL_433396|2020- ···················· ·A·················· ···················· ···················· ···················· ····················
B 9019 hCoV-19/Scotland/EDB1106/2020|EPI_ISL_433397|2020- ···················· ···················· ···················· ···················· ···················· ····················
B 9020 hCoV-19/Scotland/EDB1107/2020|EPI_ISL_433398|2020- ···················· ···················A ···················· ···················· ···················· ···················· 1
B 9021 hCoV-19/Scotland/EDB1109/2020|EPI_ISL_433399|2020- ···················· ·A·················· ···················· ···················· ···················· ····················
B 9022 hCoV-19/Scotland/CVR1034/2020|EPI_ISL_433500|2020- ···················· ·A·················· ···················· ···················· ···················· ····················
B 9023 hCoV-19/Scotland/CVR1053/2020|EPI_ISL_433501|2020- ···················· ·A·················· ···················· ···················· ···················· ····················
B 9024 hCoV-19/Scotland/CVR1056/2020|EPI_ISL_433502|2020- ···················· ···················A ···················· ···················· ···················· ···················· 1
B 9025 hCoV-19/Scotland/CVR1068/2020|EPI_ISL_433505|2020- ···················· ···················A ···················· ···················· ···················· ···················· 1
B 9026 hCoV-19/Scotland/CVR1081/2020|EPI_ISL_433507|2020- ···················· ···················· ···················· ···················· ···················· ····················
B 9027 hCoV-19/Scotland/CVR1085/2020|EPI_ISL_433508|2020- ···················· ···················· ···················· ···················· ···················· ····················
B 9028 hCoV-19/Scotland/CVR1093/2020|EPI_ISL_433509|2020- ···················· ···················A ···················· ···················· ···················· ···················· 1
B 9029 hCoV-19/Scotland/CVR1101/2020|EPI_ISL_433510|2020- ···················· ···················· ···················· ···················· ···················· ····················
B 9030 hCoV-19/Scotland/CVR1115/2020|EPI_ISL_433511|2020- ···················· ·A·················· ···················· ···················· ···················· ····················
B 9031 hCoV-19/Scotland/CVR1133/2020|EPI_ISL_433514|2020- ···················· ···················· ···················· ···················· ···················· ····················
B 9032 hCoV-19/Scotland/CVR1133b/2020|EPI_ISL_433515|2020 ···················· ···················· ···················· ···················· ···················· ····················
B 9033 hCoV-19/Scotland/CVR1140/2020|EPI_ISL_433517|2020- ···················· ···················· ···················· ···················· ···················· ····················
B 9034 hCoV-19/Scotland/CVR1142/2020|EPI_ISL_433518|2020- ···················· ···················· ···················· ···················· ···················· ····················
B 9035 hCoV-19/Scotland/CVR1145/2020|EPI_ISL_433519|2020- ···················· ···················· ···················· ···················· ···················· ····················
B 9036 hCoV-19/Scotland/CVR1159/2020|EPI_ISL_433521|2020- ···················· ···················· ···················· ···················· ···················· ····················
B 9037 hCoV-19/Scotland/CVR1204/2020|EPI_ISL_433522|2020- ···················· ·A·················· ···················· ···················· ···················· ····················
B 9038 hCoV-19/Scotland/CVR1336/2020|EPI_ISL_433545|2020- ···················· ···················A ···················· ···················· ···················· ···················· 1
B 9039 hCoV-19/Scotland/CVR1337/2020|EPI_ISL_433546|2020- ···················· ···················A ···················· ···················· ···················· ···················· 1
B 9040 hCoV-19/Scotland/CVR1343/2020|EPI_ISL_433547|2020- ···················· ···················· ···················· ···················· ···················· ····················
B 9041 hCoV-19/Scotland/CVR1345/2020|EPI_ISL_433548|2020- ···················· ···················· ···················· ···················· ···················· ····················
B 9042 hCoV-19/Scotland/CVR1351/2020|EPI_ISL_433549|2020- ···················· ···················· ···················· ···················· ···················· ····················
B 9043 hCoV-19/Scotland/CVR1352/2020|EPI_ISL_433550|2020- ···················· ·A·················· ···················· ···················· ···················· ····················
B 9044 hCoV-19/Scotland/CVR1356/2020|EPI_ISL_433551|2020- ···················· ·A·················· ···················· ···················· ···················· ····················
B 9045 hCoV-19/Scotland/CVR1358/2020|EPI_ISL_433552|2020- ···················· ·A·················· ···················· ···················· ···················· ····················
B 9046 hCoV-19/Scotland/CVR1361/2020|EPI_ISL_433554|2020- ···················· ···················· ···················· ···················· ···················· ····················
B 9047 hCoV-19/Scotland/CVR1366/2020|EPI_ISL_433557|2020- ···················· ···················· ···················· ···················· ···················· ····················
B 9048 hCoV-19/Scotland/CVR1371/2020|EPI_ISL_433559|2020- ···················· ···················· ···················· ···················· ···················· ····················
B 9049 hCoV-19/Scotland/CVR1384/2020|EPI_ISL_433563|2020- ···················· ·A·················· ···················· ···················· ···················· ····················
B 9050 hCoV-19/Sweden/20-06503/2020|EPI_ISL_434656|2020-0 ···················· ···················· ···················· ···················· ···················· ····················
B 9051 hCoV-19/Sweden/20-06813/2020|EPI_ISL_434657|2020-0 ···················· ···················· ···················· ···················· ···················· ····················
B 9052 hCoV-19/Sweden/20-06883/2020|EPI_ISL_434658|2020-0 ···················· ···················· ···················· ···················· ···················· ····················
B 9053 hCoV-19/Sweden/20-06908/2020|EPI_ISL_434659|2020-0 ···················· ···················· ···················· ···················· ···················· ····················
B 9054 hCoV-19/Sweden/20-06928/2020|EPI_ISL_434660|2020-0 ···················· ···················· ···················· ···················· ···················· ····················
B 9055 hCoV-19/Sweden/20-07010/2020|EPI_ISL_434661|2020-0 ···················· ···················· ···················· ···················· ···················· ····················
B 9056 hCoV-19/Sweden/20-07013/2020|EPI_ISL_434662|2020-0 ···················· ···················· ···················· ···················· ···················· ····················
B 9057 hCoV-19/Sweden/20-07025/2020|EPI_ISL_434663|2020-0 ···················· ···················· ···················· ···················· ···················· ····················
B 9058 hCoV-19/Sweden/20-07034/2020|EPI_ISL_434664|2020-0 ···················· ···················· ···················· ···················· ···················· ····················
B 9059 hCoV-19/Sweden/20-07042/2020|EPI_ISL_434665|2020-0 ···················· ···················· ···················· ···················· ···················· ····················
B 9060 hCoV-19/Sweden/20-07171/2020|EPI_ISL_434666|2020-0 ···················· ···················· ···················· ···················· ···················· ····················
B 9061 hCoV-19/Sweden/20-07298/2020|EPI_ISL_434667|2020-0 ···················· ···················· ···················· ···················· ···················· ····················
B 9062 hCoV-19/Sweden/20-07389/2020|EPI_ISL_434668|2020-0 ···················· ···················· ···················· ···················· ···················· ····················
B 9063 hCoV-19/Sweden/20-07390/2020|EPI_ISL_434669|2020-0 ···················· ···················· ···················· ···················· ···················· ····················
B 9064 hCoV-19/Sweden/20-07439/2020|EPI_ISL_434670|2020-0 ···················· ···················· ···················· ···················· ···················· ····················
B 9065 hCoV-19/Sweden/20-07440/2020|EPI_ISL_434671|2020-0 ···················· ···················· ···················· ···················· ···················· ····················
B 9066 hCoV-19/Sweden/20-07459/2020|EPI_ISL_434672|2020-0 ···················· ···················· ···················· ···················· ···················· ····················
B 9067 hCoV-19/Sweden/20-07480/2020|EPI_ISL_434673|2020-0 ···················· ···················· ···················· ···················· ···················· ····················
B 9068 hCoV-19/Sweden/20-07572/2020|EPI_ISL_434674|2020-0 ···················· ···················· ···················· ···················· ···················· ····················
B 9069 hCoV-19/Sweden/20-07607/2020|EPI_ISL_434675|2020-0 ···················· ···················· ···················· ···················· ···················· ····················
B 9070 hCoV-19/Sweden/20-07616/2020|EPI_ISL_434676|2020-0 ···················· ···················· ···················· ···················· ···················· ····················
B 9071 hCoV-19/env/USA/FL-UF3/2020|EPI_ISL_434677|2020-03 ···················· ···················· ···················· ···················· ···················· ····················
B 9072 hCoV-19/USA/MD-HP00015/2020|EPI_ISL_434690|2020-03 ···················· ···················· ···················· ···················· ···················· ····················
B 9074 hCoV-19/USA/TX-HMH0108/2020|EPI_ISL_434811|2020-03 ···················· ···················· ···················· ···················· ···················· ····················
B 9075 hCoV-19/USA/TX-HMH0110/2020|EPI_ISL_434812|2020-03 ···················· ···················· ···················· ···················· ···················· ····················
B 9078 hCoV-19/USA/TX-HMH0121/2020|EPI_ISL_434821|2020-03 ···················· ···················· ···················· ···················· ···················· ····················
B 9079 hCoV-19/USA/TX-HMH0122/2020|EPI_ISL_434822|2020-03 ···················· ···················· ···················· ···················· ···················· ····················
B 9080 hCoV-19/USA/TX-HMH0124/2020|EPI_ISL_434824|2020-03 ···················· ···················· ···················· ···················· ···················· ····················
B 9081 hCoV-19/USA/TX-HMH0125/2020|EPI_ISL_434825|2020-03 ···················· ···················· ···················· ···················· ···················· ····················
B 9082 hCoV-19/USA/TX-HMH0127/2020|EPI_ISL_434827|2020-03 ···················· ···················· ···················· ···················· ···················· ····················
B 9083 hCoV-19/USA/TX-HMH0128/2020|EPI_ISL_434828|2020-03 ···················· ···················· ···················· ···················· ···················· ····················
```

| | | | | | | | | |
|---|---|---|---|---|---|---|---|---|
| B | 9084 | hCoV-19/USA/TX-HMH0129/2020\|EPI_ISL_434829\|2020-03 | ···················· | ···················· | ····················· | ···················· | ···················· | ···················· |
| B | 9085 | hCoV-19/USA/TX-HMH0131/2020\|EPI_ISL_434831\|2020-03 | ···················· | ···················· | ····················· | ···················· | ···················· | ···················· |
| B | 9086 | hCoV-19/USA/TX-HMH0132/2020\|EPI_ISL_434832\|2020-03 | ···················· | ···················· | ····················· | ···················· | ···················· | ···················· |
| B | 9088 | hCoV-19/USA/TX-HMH0134/2020\|EPI_ISL_434834\|2020-03 | ···················· | ···················· | ····················· | ···················· | ···················· | ···················· |
| B | 9090 | hCoV-19/USA/TX-HMH0140/2020\|EPI_ISL_434839\|2020-03 | ···················· | ···················· | ····················· | ···················· | ···················· | ···················· |
| B | 9091 | hCoV-19/USA/TX-HMH0141/2020\|EPI_ISL_434840\|2020-03 | ···················· | ···················· | ····················· | ···················· | ···················· | ···················· |
| B | 9092 | hCoV-19/USA/TX-HMH0142/2020\|EPI_ISL_434841\|2020-03 | ···················· | ···················· | ····················· | ···················· | ···················· | ···················· |
| B | 9093 | hCoV-19/USA/TX-HMH0144/2020\|EPI_ISL_434843\|2020-03 | ···················· | ···················· | ····················· | ···················· | ···················· | ···················· |
| B | 9094 | hCoV-19/USA/TX-HMH0145/2020\|EPI_ISL_434844\|2020-03 | ···················· | ···················· | ····················· | ···················· | ···················· | ···················· |
| B | 9095 | hCoV-19/USA/TX-HMH0146/2020\|EPI_ISL_434845\|2020-03 | ···················· | ···················· | ····················· | ···················· | ···················· | ···················· |
| B | 9096 | hCoV-19/USA/TX-HMH0147/2020\|EPI_ISL_434846\|2020-03 | ···················· | ···················· | ····················· | ···················· | ···················· | ···················· |
| B | 9097 | hCoV-19/USA/TX-HMH0148/2020\|EPI_ISL_434847\|2020-03 | ···················· | ···················· | ····················· | ···················· | ···················· | ···················· |
| B | 9098 | hCoV-19/USA/TX-HMH0150/2020\|EPI_ISL_434849\|2020-03 | ···················· | ···················· | ····················· | ···················· | ···················· | ···················· |
| B | 9099 | hCoV-19/USA/TX-HMH0151/2020\|EPI_ISL_434850\|2020-03 | ···················· | ···················· | ····················· | ···················· | ···················· | ·······N············ |
| B | 9100 | hCoV-19/USA/TX-HMH0152/2020\|EPI_ISL_434851\|2020-03 | ···················· | ···················· | ····················· | ···················· | ···················· | ···················· |
| B | 9101 | hCoV-19/USA/TX-HMH0153/2020\|EPI_ISL_434852\|2020-03 | ···················· | ···················· | ····················· | ···················· | ···················· | ···················· |
| B | 9102 | hCoV-19/USA/TX-HMH0154/2020\|EPI_ISL_434853\|2020-03 | ···················· | ···················· | ····················· | ···················· | ···················· | ···················· |
| B | 9103 | hCoV-19/USA/TX-HMH0201/2020\|EPI_ISL_434876\|2020-03 | ···················· | ···················· | ····················· | ···················· | ···················· | ···················· |
| B | 9104 | hCoV-19/USA/TX-HMH0202/2020\|EPI_ISL_434877\|2020-03 | ···················· | ···················· | ····················· | ···················· | ···················· | ···················· |
| B | 9105 | hCoV-19/USA/TX-HMH0203/2020\|EPI_ISL_434878\|2020-03 | ···················· | ···················· | ····················· | ···················· | ···················· | ···················· |
| B | 9106 | hCoV-19/USA/TX-HMH0204/2020\|EPI_ISL_434879\|2020-03 | ···················· | ···················· | ····················· | ···················· | ···················· | ···················· |
| B | 9107 | hCoV-19/USA/TX-HMH0205/2020\|EPI_ISL_434880\|2020-03 | ···················· | ···················· | ····················· | ···················· | ···················· | ···················· |
| B | 9108 | hCoV-19/USA/TX-HMH0206/2020\|EPI_ISL_434881\|2020-03 | ···················· | ···················· | ····················· | ···················· | ···················· | ···················· |
| B | 9109 | hCoV-19/USA/TX-HMH0207/2020\|EPI_ISL_434882\|2020-03 | ···················· | ···················· | ····················· | ···················· | ···················· | ···················· |
| B | 9110 | hCoV-19/USA/TX-HMH0208/2020\|EPI_ISL_434883\|2020-03 | ···················· | ···················· | ····················· | ···················· | ···················· | ···················· |
| B | 9111 | hCoV-19/USA/TX-HMH0209/2020\|EPI_ISL_434884\|2020-03 | ···················· | ···················· | ····················· | ···················· | ···················· | ···················· |
| B | 9112 | hCoV-19/USA/TX-HMH0210/2020\|EPI_ISL_434885\|2020-03 | ···················· | ···················· | ····················· | ···················· | ···················· | ···················· |
| B | 9113 | hCoV-19/USA/TX-HMH0212/2020\|EPI_ISL_434887\|2020-03 | ···················· | ···················· | ····················· | ···················· | ···················· | ···················· |
| B | 9114 | hCoV-19/USA/TX-HMH0213/2020\|EPI_ISL_434888\|2020-03 | ···················· | ···················· | ····················· | ···················· | ···················· | ···················· |
| B | 9115 | hCoV-19/USA/TX-HMH0214/2020\|EPI_ISL_434889\|2020-03 | ···················· | ···················· | ····················· | ···················· | ···················· | ···················· |
| B | 9116 | hCoV-19/USA/TX-HMH0265/2020\|EPI_ISL_434894\|2020-03 | ···················· | ···················· | ····················· | ···················· | ···················· | ···················· |
| B | 9117 | hCoV-19/USA/TX-HMH0266/2020\|EPI_ISL_434895\|2020-03 | ···················· | ···················· | ····················· | ···················· | ···················· | ···················· |
| B | 9118 | hCoV-19/Latvia/011/2020\|EPI_ISL_437090\|2020-03-24 | ···················· | ···················· | ····················· | ···················· | ···················· | ···················· |
| B | 9119 | hCoV-19/Latvia/012/2020\|EPI_ISL_437091\|2020-03-23 | ···················· | ···················· | ····················· | ···················· | ···················· | ···················· |
| B | 9120 | hCoV-19/Latvia/013/2020\|EPI_ISL_437092\|2020-03-20 | ···················· | ···················· | ····················· | ···················· | ···················· | ···················· |
| B | 9121 | hCoV-19/Latvia/014/2020\|EPI_ISL_437093\|2020-03-22 | ···················· | ···················· | ····················· | ···················· | ···················· | ···················· |
| B | 9122 | hCoV-19/Latvia/015/2020\|EPI_ISL_437094\|2020-03-23 | ···················· | ···················· | ····················· | ···················· | ···················· | ···················· |
| B | 9123 | hCoV-19/Latvia/016/2020\|EPI_ISL_437095\|2020-03-23 | ···················· | ···················· | ····················· | ···················· | ···················· | ···················· |
| B | 9124 | hCoV-19/Latvia/017/2020\|EPI_ISL_437096\|2020-03-23 | ···················· | ···················· | ····················· | ···················· | ···················· | ···················· |
| B | 9125 | hCoV-19/USA/MI-MDHHS-SC20012/2020\|EPI_ISL_437099\|2 | ···················· | ···················· | ····················· | ···················· | ···················· | ···················· |
| B | 9126 | hCoV-19/England/BRIS-12505F/2020\|EPI_ISL_444006\|20 | ···················· | ···················· | ····················· | ···················· | ···················· | ···················· |
| B | 9127 | hCoV-19/England/BRIS-124C5C/2020\|EPI_ISL_444007\|20 | ···················· | ···················· | ····················· | ···················· | ···················· | ···················· |
| B | 9128 | hCoV-19/England/BRIS-125BD6/2020\|EPI_ISL_444018\|20 | ···················· | ···················· | ····················· | ···················· | ···················· | ···················· |
| B | 9129 | hCoV-19/England/BRIS-124EDE/2020\|EPI_ISL_444019\|20 | ···················· | ···················· | ····················· | ···················· | ···················· | ···················· |
| B | 9130 | hCoV-19/England/BRIS-1252E0/2020\|EPI_ISL_444020\|20 | ···················· | ···················· | ····················· | ···················· | ···················· | ···················· |
| B | 9131 | hCoV-19/England/BRIS-124B13/2020\|EPI_ISL_444021\|20 | ···················· | ···················· | ····················· | ···················· | ···················· | ···················· |
| B | 9132 | hCoV-19/USA/TX_GCID_192000003/2020\|EPI_ISL_444022\| | ···················· | ···················· | ····················· | ···················· | ···················· | ···················· |
| B | 9133 | hCoV-19/USA/CA-CZB-1089/2020\|EPI_ISL_444023\|2020-0 | ···················· | ···················· | ····················· | ···················· | ···················· | ···················· |
| B | 9134 | hCoV-19/USA/CA-CZB-1090/2020\|EPI_ISL_444024\|2020-0 | ···················· | ···················· | ····················· | ···················· | ···················· | ···················· |
| B | 9135 | hCoV-19/USA/CA-CZB-1091/2020\|EPI_ISL_444025\|2020-0 | ···················· | ···················· | ····················· | ···················· | ···················· | ···················· |
| B | 9136 | hCoV-19/USA/CA-CZB-1092/2020\|EPI_ISL_444026\|2020-0 | ···················· | ···················· | ····················· | ···················· | ···················· | ···················· |
| B | 9137 | hCoV-19/USA/CA-CZB-1093/2020\|EPI_ISL_444051\|2020-0 | ···················· | ···················· | ····················· | ···················· | ···················· | ···················· |
| B | 9138 | hCoV-19/USA/CA-CZB-1094/2020\|EPI_ISL_444052\|2020-0 | ···················· | ···················· | ····················· | ···················· | ···················· | ···················· |
| B | 9139 | hCoV-19/USA/CA-CZB-1095/2020\|EPI_ISL_444053\|2020-0 | ···················· | ···················· | ····················· | ···················· | ···················· | ···················· |
| B | 9140 | hCoV-19/USA/CA-CZB-1096/2020\|EPI_ISL_444054\|2020-0 | ···················· | ···················· | ····················· | ···················· | ···················· | ···················· |
| B | 9141 | hCoV-19/USA/CA-CZB-1097/2020\|EPI_ISL_444055\|2020-0 | ···················· | ···················· | ····················· | ···················· | ···················· | ···················· |
| B | 9142 | hCoV-19/USA/CA-CZB-1099/2020\|EPI_ISL_444057\|2020-0 | ···················· | ···················· | ····················· | ···················· | ···················· | ···················· |
| B | 9143 | hCoV-19/USA/CA-CZB-1100/2020\|EPI_ISL_444058\|2020-0 | ···················· | ···················· | ····················· | ···················· | ···················· | ···················· |
| B | 9144 | hCoV-19/USA/CA-CZB-1101/2020\|EPI_ISL_444059\|2020-0 | ···················· | ···················· | ····················· | ···················· | ···················· | ···················· |
| B | 9145 | hCoV-19/England/LOND-D50E6/2020\|EPI_ISL_444201\|202 | ···················· | ···················· | ····················· | ···················· | ···················· | ···················· |
| B | 9146 | hCoV-19/England/LOND-D50F5/2020\|EPI_ISL_444202\|202 | ···················· | ···················· | ····················· | ···················· | ···················· | ···················· |
| B | 9147 | hCoV-19/England/LOND-D512F/2020\|EPI_ISL_444203\|202 | ···················· | ···················· | ····················· | ···················· | ···················· | ···················· |
| B | 9148 | hCoV-19/England/LOND-D531A/2020\|EPI_ISL_444210\|202 | ···················· | ···················· | ····················· | ···················· | ···················· | ···················· |
| B | 9149 | hCoV-19/England/LOND-D5329/2020\|EPI_ISL_444211\|202 | ···················· | ···················· | ····················· | ···················· | ···················· | ···················· |
| B | 9150 | hCoV-19/England/LOND-D5338/2020\|EPI_ISL_444212\|202 | ···················· | ···················· | ····················· | ···················· | ···················· | ···················· |
| B | 9151 | hCoV-19/England/LOND-D5365/2020\|EPI_ISL_444213\|202 | ···················· | ···················· | ····················· | ···················· | ···················· | ···················· |
| B | 9152 | hCoV-19/England/LOND-D5383/2020\|EPI_ISL_444214\|202 | ···················· | ···················· | ····················· | ···················· | ···················· | ···················· |
| B | 9153 | hCoV-19/Chile/RM-ISPCH-25/2020\|EPI_ISL_445304\|2020 | ···················· | ···················· | ····················· | ···················· | ···················· | ···················· |
| B | 9154 | hCoV-19/Chile/RM-ISPCH-26/2020\|EPI_ISL_445305\|2020 | ···················· | ···················· | ····················· | ···················· | ···················· | ···················· |
| B | 9155 | hCoV-19/Chile/RM-ISPCH-27/2020\|EPI_ISL_445306\|2020 | ···················· | ···················· | ····················· | ···················· | ···················· | ···················· |
| B | 9156 | hCoV-19/Chile/RM-ISPCH-28/2020\|EPI_ISL_445307\|2020 | ···················· | ···················· | ····················· | ···················· | ···················· | ···················· |
| B | 9157 | hCoV-19/Chile/RM-ISPCH-30/2020\|EPI_ISL_445309\|2020 | ···················· | ···················· | ····················· | ···················· | ···················· | ···················· |
| B | 9158 | hCoV-19/Chile/RM-ISPCH-31/2020\|EPI_ISL_445310\|2020 | ···················· | ···················· | ····················· | ···················· | ···················· | ···················· |
| B | 9159 | hCoV-19/Chile/RM-ISPCH-32/2020\|EPI_ISL_445311\|2020 | ···················· | ···················· | ····················· | ···················· | ···················· | ···················· |
| B | 9160 | hCoV-19/Chile/RM-ISPCH-33/2020\|EPI_ISL_445312\|2020 | ···················· | ···················· | ····················· | ···················· | ···················· | ···················· |
| B | 9161 | hCoV-19/Chile/RM-ISPCH-34/2020\|EPI_ISL_445313\|2020 | ···················· | ···················· | ····················· | ···················· | ···················· | ···················· |
| B | 9162 | hCoV-19/Chile/RM-ISPCH-35/2020\|EPI_ISL_445314\|2020 | ···················· | ···················· | ····················· | ···················· | ···················· | ···················· |
| B | 9163 | hCoV-19/Chile/RM-ISPCH-36/2020\|EPI_ISL_445315\|2020 | ···················· | ···················· | ····················· | ···················· | ···················· | ···················· |

| | | | | | | | | | |
|---|---|---|---|---|---|---|---|---|---|
| B | 9164 | hCoV-19/Chile/RM-ISPCH-37/2020\|EPI_ISL_445316\|2020 | ···················· | ···················· | ···················· | ···················· | ···················· | ···················· | |
| B | 9165 | hCoV-19/Chile/RM-ISPCH-38/2020\|EPI_ISL_445317\|2020 | ···················· | ···················· | ···················· | ···················· | ···················· | ···················· | |
| B | 9166 | hCoV-19/Chile/RM-ISPCH-39/2020\|EPI_ISL_445318\|2020 | ···················· | ···················· | ···················· | ···················· | ···················· | ···················· | |
| B | 9167 | hCoV-19/Chile/RM-ISPCH-41/2020\|EPI_ISL_445320\|2020 | ···················· | ···················· | ···················· | ···················· | ···················· | ···················· | |
| B | 9168 | hCoV-19/Chile/RM-ISPCH-43/2020\|EPI_ISL_445322\|2020 | ···················· | ···················· | ···················· | ···················· | ···················· | ···················· | |
| B | 9169 | hCoV-19/Chile/RM-ISPCH-44/2020\|EPI_ISL_445323\|2020 | ···················· | ···················· | ···················· | ···················· | ···················· | ···················· | |
| B | 9170 | hCoV-19/Chile/AR-ISPCH-14/2020\|EPI_ISL_445348\|2020 | ···················· | ···················· | ···················· | ···················· | ···················· | ···················· | |
| B | 9171 | hCoV-19/Chile/RM-ISPCH-52/2020\|EPI_ISL_445349\|2020 | ···················· | ···················· | ···················· | ···················· | ···················· | ···················· | |
| B | 9172 | hCoV-19/Chile/RM-ISPCH-54/2020\|EPI_ISL_445350\|2020 | ···················· | ···················· | ···················· | ···················· | ···················· | ···················· | |
| B | 9173 | hCoV-19/Chile/RM-ISPCH-55/2020\|EPI_ISL_445351\|2020 | ···················· | ···················· | ···················· | ···················· | ···················· | ···················· | |
| B | 9174 | hCoV-19/Chile/RM-ISPCH-56/2020\|EPI_ISL_445352\|2020 | ···················· | ···················· | ···················· | ···················· | ···················· | ···················· | |
| B | 9175 | hCoV-19/Chile/RM-ISPCH-57/2020\|EPI_ISL_445353\|2020 | ···················· | ···················· | ···················· | ···················· | ···················· | ···················· | |
| B | 9176 | hCoV-19/Chile/RM-ISPCH-59/2020\|EPI_ISL_445355\|2020 | ···················· | ···················· | ···················· | ···················· | ···················· | ···················· | |
| B | 9177 | hCoV-19/Chile/RM-ISPCH-60/2020\|EPI_ISL_445356\|2020 | ···················· | ···················· | ···················· | ···················· | ···················· | ···················· | |
| B | 9178 | hCoV-19/Chile/RM-ISPCH-62/2020\|EPI_ISL_445358\|2020 | ···················· | ···················· | ···················· | ···················· | ···················· | ···················· | |
| B | 9179 | hCoV-19/Chile/RM-ISPCH-64/2020\|EPI_ISL_445360\|2020 | ···················· | ···················· | ···················· | ···················· | ···················· | ···················· | |
| B | 9180 | hCoV-19/Chile/RM-ISPCH-65/2020\|EPI_ISL_445361\|2020 | ···················· | ···················· | ···················· | ···················· | ···················· | ···················· | |
| B | 9181 | hCoV-19/Chile/RM-ISPCH-68/2020\|EPI_ISL_445364\|2020 | ···················· | ···················· | ···················· | ···················· | ···················· | ···················· | |
| B | 9182 | hCoV-19/Chile/RM-ISPCH-69/2020\|EPI_ISL_445365\|2020 | ···················· | ···················· | ···················· | ···················· | ···················· | ···················· | |
| B | 9183 | hCoV-19/Chile/RM-ISPCH-70/2020\|EPI_ISL_445366\|2020 | ···················· | ···················· | ···················· | ···················· | ···················· | ···················· | |
| B | 9184 | hCoV-19/Chile/RM-ISPCH-71/2020\|EPI_ISL_445367\|2020 | ···················· | ···················· | ···················· | ···················· | ···················· | ···················· | |
| B | 9185 | hCoV-19/Chile/RM-ISPCH-72/2020\|EPI_ISL_445368\|2020 | ···················· | ···················· | ···················· | ···················· | ···················· | ···················· | |
| B | 9186 | hCoV-19/Chile/RM-ISPCH-75/2020\|EPI_ISL_445371\|2020 | ···················· | ···················· | ···················· | ···················· | ···················· | ···················· | |
| B | 9187 | hCoV-19/Chile/MA-ISPCH-17/2020\|EPI_ISL_445372\|2020 | ···················· | ···················· | ···················· | ···················· | ···················· | ···················· | |
| B | 9188 | hCoV-19/Chile/RM-ISPCH-76/2020\|EPI_ISL_445373\|2020 | ···················· | ···················· | ···················· | ···················· | ···················· | ···················· | |
| B | 9189 | hCoV-19/Chile/RM-ISPCH-78/2020\|EPI_ISL_445375\|2020 | ···················· | ···················· | ···················· | ···················· | ···················· | ···················· | |
| B | 9190 | hCoV-19/Chile/RM-ISPCH-79/2020\|EPI_ISL_445376\|2020 | ···················· | ···················· | ···················· | ···················· | ···················· | ···················· | |
| B | 9191 | hCoV-19/Chile/RM-ISPCH-80/2020\|EPI_ISL_445377\|2020 | ···················· | ···················· | ···················· | ···················· | ···················· | ···················· | |
| B | 9192 | hCoV-19/Chile/BI-ISPCH-4/2020\|EPI_ISL_445378\|2020- | ···················· | ···················· | ···················· | ···················· | ···················· | ···················· | |
| B | 9193 | hCoV-19/Wales/PHWC-289F8/2020\|EPI_ISL_445385\|2020- | ···················· | ·A·················· | ···················· | ···················· | ···················· | ···················· | |
| B | 9194 | hCoV-19/Wales/PHWC-28A13/2020\|EPI_ISL_445387\|2020- | ···················· | ···················· | ···················· | ···················· | ···················· | ···················· | |
| B | 9195 | hCoV-19/Wales/PHWC-28A22/2020\|EPI_ISL_445388\|2020- | ···················· | ···················· | ···················· | ···················· | ···················· | ···················· | |
| B | 9196 | hCoV-19/Wales/PHWC-29265/2020\|EPI_ISL_445500\|2020- | ···················· | ···················· | ···················· | ···················· | ···················· | ···················· | |
| B | 9197 | hCoV-19/Wales/PHWC-2956C/2020\|EPI_ISL_445531\|2020- | ···················· | ···················· | ···················· | ···················· | ···················· | ···················· | |
| B | 9198 | hCoV-19/Wales/PHWC-2A246/2020\|EPI_ISL_445541\|2020- | ···················· | ···················· | ···················· | ···················· | ···················· | ···················· | |
| B | 9199 | hCoV-19/Wales/PHWC-2A282/2020\|EPI_ISL_445545\|2020- | ···················· | ···················· | ···················· | ···················· | ···················· | ···················· | |
| B | 9200 | hCoV-19/Wales/PHWC-32348/2020\|EPI_ISL_446636\|2020- | ···················· | ···················· | ···················· | ···················· | ···················· | ···················· | |
| B | 9201 | hCoV-19/Wales/PHWC-32375/2020\|EPI_ISL_446638\|2020- | ···················· | ···················· | ···················· | ···················· | ···················· | ···················· | |
| B | 9202 | hCoV-19/Wales/PHWC-32393/2020\|EPI_ISL_446639\|2020- | ···················· | ···················· | ···················· | ···················· | ···················· | ···················· | |
| B | 9203 | hCoV-19/Wales/PHWC-323EE/2020\|EPI_ISL_446643\|2020- | ···················· | ···················· | ···················· | ···················· | ···················· | ···················· | |
| B | 9204 | hCoV-19/Wales/PHWC-323FD/2020\|EPI_ISL_446644\|2020- | ···················· | ···················· | ···················· | ···················· | ···················· | ···················· | |
| B | 9205 | hCoV-19/Wales/PHWC-32418/2020\|EPI_ISL_446646\|2020- | ···················· | ···················· | ···················· | ···················· | ···················· | ···················· | |
| B | 9206 | hCoV-19/Wales/PHWC-32427/2020\|EPI_ISL_446647\|2020- | ···················· | ···················· | ···················· | ···················· | ···················· | ···················· | |
| B | 9207 | hCoV-19/Wales/PHWC-32472/2020\|EPI_ISL_446649\|2020- | ···················· | ···················· | ···················· | ···················· | ···················· | ···················· | |
| B | 9208 | hCoV-19/Wales/PHWC-32CF2/2020\|EPI_ISL_446679\|2020- | ···················· | ···················· | ···················· | ···················· | NNN················· | ···················· | 1 |
| B | 9209 | hCoV-19/Wales/PHWC-32DD1/2020\|EPI_ISL_446691\|2020- | ···················· | ···················· | ···················· | ···················· | ···················· | ···················· | |
| B | 9210 | hCoV-19/Wales/PHWC-32E65/2020\|EPI_ISL_446698\|2020- | ···················· | ···················· | ···················· | ···················· | ···················· | ···················· | |
| B | 9212 | hCoV-19/Wales/PHWC-33C1F/2020\|EPI_ISL_446802\|2020- | ···················· | ···················· | ···················· | ···················· | ···················· | ···················· | |
| B | 9213 | hCoV-19/Wales/PHWC-33CD3/2020\|EPI_ISL_446811\|2020- | ···················· | ···················· | ···················· | ···················· | ···················· | ···················· | |
| B | 9214 | hCoV-19/Wales/PHWC-33DD0/2020\|EPI_ISL_446823\|2020- | ···················· | ···················· | ···················· | ···················· | ···················· | ···················· | |
| B | 9215 | hCoV-19/Wales/PHWC-3406D/2020\|EPI_ISL_446862\|2020- | ···················· | ···················· | ···················· | ···················· | ···················· | ···················· | |
| B | 9216 | hCoV-19/Wales/PHWC-3411F/2020\|EPI_ISL_446873\|2020- | ···················· | ···················· | ···················· | ···················· | ···················· | ···················· | |
| B | 9217 | hCoV-19/Wales/PHWC-276C5/2020\|EPI_ISL_432200\|2020- | ···················· | ···A················ | ···················· | ···················· | ···················· | ···················· | |
| B | 9218 | hCoV-19/Wales/PHWC-2A0E2/2020\|EPI_ISL_432201\|2020- | ···················· | ···················· | ···················· | ···················· | ···················· | ···················· | |
| B | 9219 | hCoV-19/Wales/PHWC-29E8F/2020\|EPI_ISL_432202\|2020- | ···················· | ···················· | ···················· | ···················· | ···················· | ···················· | |
| B | 9220 | hCoV-19/Wales/PHWC-29E52/2020\|EPI_ISL_432203\|2020- | ···················· | ···················· | ···················· | ···················· | ···················· | ···················· | |
| B | 9221 | hCoV-19/Wales/PHWC-2A10D/2020\|EPI_ISL_432204\|2020- | ···················· | ···················· | ···················· | ···················· | ···················· | ···················· | |
| B | 9222 | hCoV-19/Wales/PHWC-29E34/2020\|EPI_ISL_432205\|2020- | ···················· | ···················· | ···················· | ···················· | ···················· | ···················· | |
| B | 9223 | hCoV-19/Wales/PHWC-27698/2020\|EPI_ISL_432206\|2020- | ···················· | ···················· | ···················· | ···················· | ···················· | ···················· | |
| B | 9224 | hCoV-19/Wales/PHWC-2530C/2020\|EPI_ISL_432207\|2020- | ···················· | ···················· | ···················· | ···················· | ···················· | ···················· | |
| B | 9225 | hCoV-19/Wales/PHWC-278A1/2020\|EPI_ISL_432208\|2020- | ···················· | ···················· | ···················· | ···················· | ···················· | ···················· | |
| B | 9226 | hCoV-19/Wales/PHWC-27A41/2020\|EPI_ISL_432209\|2020- | ···················· | ···A················ | ···················· | ···················· | ···················· | ···················· | |
| B | 9227 | hCoV-19/Wales/PHWC-277B3/2020\|EPI_ISL_432210\|2020- | ···················· | ···················· | ···················· | ···················· | ···················· | ···················· | |
| B | 9228 | hCoV-19/Wales/PHWC-2764D/2020\|EPI_ISL_432211\|2020- | ···················· | ···················· | ···················· | ···················· | ···················· | ···················· | |
| B | 9229 | hCoV-19/Wales/PHWC-27944/2020\|EPI_ISL_432212\|2020- | ···················· | ···················· | ···················· | ···················· | ···················· | ···················· | |
| B | 9230 | hCoV-19/Wales/PHWC-2AF5E/2020\|EPI_ISL_432213\|2020- | ···················· | ···················· | ···················· | ···················· | ···················· | ···················· | |
| B | 9231 | hCoV-19/Wales/PHWC-2A0B5/2020\|EPI_ISL_432230\|2020- | ···················· | ···················· | ···················· | ···················· | ···················· | ···················· | |
| B | 9232 | hCoV-19/Wales/PHWC-2532A/2020\|EPI_ISL_432231\|2020- | ···················· | ···················· | ···················· | ···················· | ···················· | ···················· | |
| B | 9233 | hCoV-19/Wales/PHWC-275D7/2020\|EPI_ISL_432233\|2020- | ···················· | ···················· | ···················· | ···················· | ···················· | ···················· | |
| B | 9234 | hCoV-19/Wales/PHWC-2A097/2020\|EPI_ISL_432234\|2020- | ···················· | ···················· | ···················· | ···················· | ···················· | ···················· | |
| B | 9235 | hCoV-19/Wales/PHWC-272FE/2020\|EPI_ISL_432235\|2020- | ···················· | ···················· | ···················· | ···················· | ···················· | ···················· | |
| B | 9236 | hCoV-19/Scotland/CVR1261/2020\|EPI_ISL_433525\|2020- | ···················· | ···················· | ···················· | ···················· | ···················· | ···················· | |
| B | 9237 | hCoV-19/Scotland/CVR1267/2020\|EPI_ISL_433527\|2020- | ···················· | ···················· | ···················· | ···················· | ···················· | ···················· | |
| B | 9238 | hCoV-19/Scotland/CVR1271/2020\|EPI_ISL_433529\|2020- | ···················· | ·A·················· | ···················· | ···················· | ···················· | ···················· | |
| B | 9239 | hCoV-19/Scotland/CVR1305/2020\|EPI_ISL_433534\|2020- | ···················· | ···················A | ···················· | ···················· | ···················· | ···················· | 1 |
| B | 9240 | hCoV-19/Scotland/CVR1308/2020\|EPI_ISL_433535\|2020- | ···················· | ···················· | ···················· | ···················· | ···················· | ···················· | |
| B | 9241 | hCoV-19/Scotland/CVR1309/2020\|EPI_ISL_433536\|2020- | ···················· | ···················· | ···················· | ···················· | ···················· | ···················· | |
| B | 9242 | hCoV-19/Scotland/CVR1311/2020\|EPI_ISL_433537\|2020- | ···················· | ···················· | ···················· | ···················· | ···················· | ···················· | |

| | | | | | | | | |
|---|---|---|---|---|---|---|---|---|
| B | 9243 hCoV-19/Scotland/CVR1312/2020\|EPI_ISL_433538\|2020- | .................... | .................... | ..................... | .................... | .................... | .................... | |
| B | 9244 hCoV-19/Scotland/CVR1314/2020\|EPI_ISL_433539\|2020- | .................... | .A.................. | ..................... | .................... | .................... | .................... | |
| B | 9245 hCoV-19/Scotland/CVR1325/2020\|EPI_ISL_433541\|2020- | .................... | .................... | ..................... | .................... | .................... | .................... | |
| B | 9246 hCoV-19/Scotland/CVR1329/2020\|EPI_ISL_433542\|2020- | .................... | ...................A | ..................... | .................... | .................... | .................... | 1 |
| B | 9247 hCoV-19/Scotland/CVR1331/2020\|EPI_ISL_433543\|2020- | .................... | ...................A | ..................... | .................... | .................... | .................... | 1 |
| B | 9248 hCoV-19/Scotland/CVR1334/2020\|EPI_ISL_433544\|2020- | .................... | .................... | ..................... | .................... | .................... | .................... | |
| B | 9249 hCoV-19/Scotland/CVR1373/2020\|EPI_ISL_433560\|2020- | .................... | .................... | ..................... | .................... | .................... | .................... | |
| B | 9250 hCoV-19/Scotland/CVR1377/2020\|EPI_ISL_433561\|2020- | .................... | .................... | ..................... | .................... | .................... | .................... | |
| B | 9251 hCoV-19/Scotland/CVR1379/2020\|EPI_ISL_433562\|2020- | .................... | .A.................. | ..................... | .................... | .................... | .................... | |
| B | 9252 hCoV-19/Scotland/CVR1387/2020\|EPI_ISL_433565\|2020- | .................... | .A.................. | ..................... | .................... | .................... | .................... | |
| B | 9253 hCoV-19/Scotland/CVR1393/2020\|EPI_ISL_433566\|2020- | .................... | .................... | ..................... | .................... | .................... | .................... | |
| B | 9254 hCoV-19/USA/TX-HMH095/2020\|EPI_ISL_434800\|2020-03- | .................... | .................... | ..................... | .................... | .................... | .................... | |
| B | 9255 hCoV-19/USA/TX-HMH096/2020\|EPI_ISL_434801\|2020-03- | .................... | .................... | ..................... | .................... | .................... | .................... | |
| B | 9257 hCoV-19/USA/TX-HMH098/2020\|EPI_ISL_434803\|2020-03- | .................... | .................... | ..................... | .................... | .................... | .................... | |
| B | 9258 hCoV-19/USA/TX-HMH0100/2020\|EPI_ISL_434805\|2020-03 | .................... | .................... | ..................... | .................... | .................... | .................... | |
| B | 9259 hCoV-19/USA/TX-HMH0102/2020\|EPI_ISL_434807\|2020-03 | .................... | .................... | ..................... | .................... | .................... | .................... | |
| B | 9261 hCoV-19/USA/TX-HMH0155/2020\|EPI_ISL_434854\|2020-03 | .................... | .................... | ..................... | .................... | .................... | .................... | |
| B | 9262 hCoV-19/USA/TX-HMH0156/2020\|EPI_ISL_434855\|2020-03 | .................... | .................... | ..................... | .................... | .................... | .................... | |
| B | 9263 hCoV-19/USA/TX-HMH0157/2020\|EPI_ISL_434856\|2020-03 | .................... | .................... | ..................... | .................... | .................... | .................... | |
| B | 9264 hCoV-19/USA/TX-HMH0158/2020\|EPI_ISL_434857\|2020-03 | .................... | .................... | ..................... | .................... | .................... | .................... | |
| B | 9265 hCoV-19/USA/TX-HMH0159/2020\|EPI_ISL_434858\|2020-03 | .................... | .................... | ..................... | .................... | .................... | .................... | |
| B | 9266 hCoV-19/USA/TX-HMH0160/2020\|EPI_ISL_434859\|2020-03 | .................... | .................... | ..................... | .................... | .................... | .................... | |
| B | 9267 hCoV-19/USA/TX-HMH0182/2020\|EPI_ISL_434860\|2020-03 | .................... | .................... | ..................... | .................... | .................... | .................... | |
| B | 9268 hCoV-19/USA/TX-HMH0183/2020\|EPI_ISL_434861\|2020-03 | .................... | .................... | ..................... | .................... | .................... | .................... | |
| B | 9269 hCoV-19/USA/TX-HMH0185/2020\|EPI_ISL_434863\|2020-03 | .................... | .................... | ..................... | .................... | .................... | .................... | |
| B | 9270 hCoV-19/USA/TX-HMH0186/2020\|EPI_ISL_434864\|2020-03 | .................... | .................... | ..................... | .................... | .................... | .................... | |
| B | 9271 hCoV-19/USA/TX-HMH0189/2020\|EPI_ISL_434865\|2020-03 | .................... | .................... | ..................... | .................... | .................... | .................... | |
| B | 9272 hCoV-19/USA/TX-HMH0191/2020\|EPI_ISL_434866\|2020-03 | .................... | .................... | ..................... | .................... | .................... | .................... | |
| B | 9273 hCoV-19/USA/TX-HMH0192/2020\|EPI_ISL_434867\|2020-03 | .................... | .................... | ..................... | .................... | .................... | .................... | |
| B | 9274 hCoV-19/USA/TX-HMH0193/2020\|EPI_ISL_434868\|2020-03 | .................... | .................... | ..................... | .................... | .................... | .................... | |
| B | 9275 hCoV-19/USA/TX-HMH0195/2020\|EPI_ISL_434870\|2020-03 | .................... | .................... | ..................... | .................... | .................... | .................... | |
| B | 9276 hCoV-19/USA/TX-HMH0196/2020\|EPI_ISL_434871\|2020-03 | .................... | .................... | ..................... | .................... | .................... | .................... | |
| B | 9277 hCoV-19/USA/TX-HMH0198/2020\|EPI_ISL_434873\|2020-03 | .................... | .................... | ..................... | .................... | .................... | .................... | |
| B | 9278 hCoV-19/USA/TX-HMH0200/2020\|EPI_ISL_434875\|2020-03 | .................... | .................... | ..................... | .................... | .................... | .................... | |
| B | 9279 hCoV-19/USA/TX-HMH0230/2020\|EPI_ISL_434890\|2020-03 | .................... | .................... | ..................... | .................... | .................... | .................... | |
| B | 9280 hCoV-19/USA/TX-HMH0234/2020\|EPI_ISL_434891\|2020-03 | .................... | .................... | ..................... | .................... | .................... | .................... | |
| B | 9281 hCoV-19/USA/TX-HMH0263/2020\|EPI_ISL_434892\|2020-03 | .................... | .................... | ..................... | .................... | .................... | .................... | |
| B | 9282 hCoV-19/USA/TX-HMH0264/2020\|EPI_ISL_434893\|2020-03 | .................... | .................... | ..................... | .................... | .................... | .................... | |
| B | 9283 hCoV-19/USA/TX-HMH0267/2020\|EPI_ISL_434896\|2020-03 | .................... | .................... | ..................... | .................... | .................... | .................... | |
| B | 9284 hCoV-19/USA/TX-HMH0268/2020\|EPI_ISL_434897\|2020-03 | .................... | .................... | ..................... | .................... | .................... | .................... | |
| B | 9285 hCoV-19/Denmark/ALAB-SSI-635/2020\|EPI_ISL_437011\|2 | .................... | .A.................. | ..................... | .................... | .................... | .................... | |
| B | 9286 hCoV-19/Denmark/ALAB-SSI-639/2020\|EPI_ISL_437013\|2 | .................... | .................... | ..................... | .................... | .................... | .................... | |
| B | 9287 hCoV-19/Denmark/ALAB-SSI-640/2020\|EPI_ISL_437014\|2 | .................... | .................... | ..................... | .................... | .................... | .................... | |
| B | 9288 hCoV-19/Denmark/ALAB-SSI-643/2020\|EPI_ISL_437015\|2 | .................... | .................... | ..................... | .................... | .................... | .................... | |
| B | 9289 hCoV-19/Denmark/ALAB-SSI-644/2020\|EPI_ISL_437016\|2 | .................... | .................... | ..................... | .................... | .................... | .................... | |
| B | 9290 hCoV-19/Denmark/ALAB-SSI-645/2020\|EPI_ISL_437017\|2 | .................... | .................... | ..................... | .................... | .................... | .................... | |
| B | 9291 hCoV-19/Denmark/ALAB-SSI-646/2020\|EPI_ISL_437018\|2 | .................... | .................... | ..................... | .................... | .................... | .................... | |
| B | 9292 hCoV-19/Denmark/ALAB-SSI-648/2020\|EPI_ISL_437019\|2 | .................... | .................... | ..................... | .................... | .................... | .................... | |
| B | 9293 hCoV-19/Denmark/ALAB-SSI-649/2020\|EPI_ISL_437020\|2 | .................... | .................... | ..................... | .................... | .................... | .................... | |
| B | 9294 hCoV-19/Denmark/ALAB-SSI-650/2020\|EPI_ISL_437021\|2 | .................... | .................... | ..................... | .................... | .................... | .................... | |
| B | 9295 hCoV-19/Denmark/ALAB-SSI-652/2020\|EPI_ISL_437023\|2 | .................... | .................... | ..................... | .................... | .................... | .................... | |
| B | 9296 hCoV-19/Denmark/ALAB-SSI-653/2020\|EPI_ISL_437024\|2 | .................... | .................... | ..................... | .................... | .................... | .................... | |
| B | 9297 hCoV-19/Denmark/ALAB-SSI-654/2020\|EPI_ISL_437025\|2 | .................... | .................... | ..................... | .................... | .................... | .................... | |
| B | 9298 hCoV-19/Denmark/ALAB-SSI-659/2020\|EPI_ISL_437026\|2 | .................... | .................... | ..................... | .................... | .................... | .................... | |
| B | 9299 hCoV-19/Denmark/ALAB-SSI-660/2020\|EPI_ISL_437027\|2 | .................... | .................... | ..................... | .................... | .................... | .................... | |
| B | 9300 hCoV-19/Denmark/ALAB-SSI-661/2020\|EPI_ISL_437028\|2 | .................... | .................... | ..................... | .................... | .................... | .................... | |
| B | 9301 hCoV-19/Denmark/ALAB-SSI-662/2020\|EPI_ISL_437029\|2 | .................... | .................... | ..................... | .................... | .................... | .................... | |
| B | 9302 hCoV-19/Denmark/ALAB-SSI-670/2020\|EPI_ISL_437030\|2 | .................... | .................... | ..................... | .................... | .................... | .................... | |
| B | 9303 hCoV-19/Denmark/ALAB-SSI-671/2020\|EPI_ISL_437031\|2 | .................... | .................... | ..................... | .................... | .................... | .................... | |
| B | 9304 hCoV-19/Denmark/ALAB-SSI-672/2020\|EPI_ISL_437032\|2 | .................... | .................... | ..................... | .................... | .................... | .................... | |
| B | 9305 hCoV-19/Denmark/ALAB-SSI-673/2020\|EPI_ISL_437033\|2 | .................... | .................... | ..................... | .................... | .................... | .................... | |
| B | 9306 hCoV-19/Denmark/ALAB-SSI-675/2020\|EPI_ISL_437035\|2 | .................... | .................... | ..................... | .................... | .................... | .................... | |
| B | 9307 hCoV-19/Denmark/ALAB-SSI-676/2020\|EPI_ISL_437036\|2 | .................... | .................... | ..................... | .................... | .................... | .................... | |
| B | 9308 hCoV-19/Denmark/ALAB-SSI-679/2020\|EPI_ISL_437037\|2 | .................... | .................... | ..................... | .................... | .................... | .................... | |
| B | 9309 hCoV-19/Denmark/ALAB-SSI-688/2020\|EPI_ISL_437038\|2 | .................... | .................... | ..................... | .................... | .................... | .................... | |
| B | 9310 hCoV-19/Denmark/ALAB-SSI-689/2020\|EPI_ISL_437039\|2 | .................... | .................... | ..................... | .................... | .................... | .................... | |
| B | 9311 hCoV-19/Denmark/ALAB-SSI-690/2020\|EPI_ISL_437040\|2 | .................... | .................... | ..................... | .................... | .................... | .................... | |
| B | 9312 hCoV-19/Denmark/ALAB-SSI-691/2020\|EPI_ISL_437041\|2 | .................... | .................... | ..................... | .................... | .................... | .................... | |
| B | 9313 hCoV-19/Denmark/ALAB-SSI-692/2020\|EPI_ISL_437042\|2 | .................... | .................... | ..................... | .................... | .................... | .................... | |
| B | 9314 hCoV-19/USA/CA-CZB-1044/2020\|EPI_ISL_437044\|2020-0 | .................... | .................... | ..................... | .................... | .................... | .................... | |
| B | 9315 hCoV-19/USA/CA-CZB-1045/2020\|EPI_ISL_437045\|2020-0 | .................... | .................... | ..................... | .................... | .................... | .................... | |
| B | 9316 hCoV-19/USA/CA-CZB-1046/2020\|EPI_ISL_437046\|2020-0 | .................... | .................... | ..................... | .................... | .................... | .................... | |
| B | 9317 hCoV-19/USA/CA-CZB-1047/2020\|EPI_ISL_437047\|2020-0 | .................... | .................... | ..................... | .................... | .................... | .................... | |
| B | 9318 hCoV-19/USA/CA-CZB-1048/2020\|EPI_ISL_437048\|2020-0 | .................... | .................... | ..................... | .................... | .................... | .................... | |
| B | 9319 hCoV-19/USA/CA-CZB-1049/2020\|EPI_ISL_437049\|2020-0 | .................... | .................... | ..................... | .................... | .................... | .................... | |
| B | 9320 hCoV-19/USA/CA-CZB-1050/2020\|EPI_ISL_437050\|2020-0 | .................... | .................... | ..................... | .................... | .................... | .................... | |
| B | 9321 hCoV-19/USA/CA-CZB-1051/2020\|EPI_ISL_437051\|2020-0 | .................... | .................... | ..................... | .................... | .................... | .................... | |
| B | 9322 hCoV-19/USA/CA-CZB-1053/2020\|EPI_ISL_437053\|2020-0 | .................... | .................... | ..................... | .................... | .................... | .................... | |

| | | | | | | | | |
|---|---|---|---|---|---|---|---|---|
| B | 9323 | hCoV-19/USA/CA-CZB-1054/2020\|EPI_ISL_437054\|2020-0 | ···················· | ···················· | ···················· | ···················· | ···················· | ···················· |
| B | 9324 | hCoV-19/USA/CA-CZB-1055/2020\|EPI_ISL_437055\|2020-0 | ···················· | ···················· | ···················· | ···················· | ···················· | ···················· |
| B | 9325 | hCoV-19/USA/CA-CZB-1056/2020\|EPI_ISL_437056\|2020-0 | ···················· | ···················· | ···················· | ···················· | ···················· | ···················· |
| B | 9326 | hCoV-19/USA/CA-CZB-1057/2020\|EPI_ISL_437057\|2020-0 | ···················· | ···················· | ···················· | ···················· | ···················· | ···················· |
| B | 9327 | hCoV-19/USA/CA-CZB-1059/2020\|EPI_ISL_437059\|2020-0 | ···················· | ···················· | ···················· | ···················· | ···················· | ···················· |
| B | 9328 | hCoV-19/USA/CA-CZB-1060/2020\|EPI_ISL_437060\|2020-0 | ···················· | ···················· | ···················· | ···················· | ···················· | ···················· |
| B | 9329 | hCoV-19/USA/CA-CZB-1061/2020\|EPI_ISL_437061\|2020-0 | ···················· | ···················· | ···················· | ···················· | ···················· | ···················· |
| B | 9330 | hCoV-19/USA/CA-CZB-1063/2020\|EPI_ISL_437063\|2020-0 | ···················· | ···················· | ···················· | ···················· | ···················· | ···················· |
| B | 9331 | hCoV-19/USA/CA-CZB-1064/2020\|EPI_ISL_437064\|2020-0 | ···················· | ···················· | ···················· | ···················· | ···················· | ···················· |
| B | 9332 | hCoV-19/USA/CA-CZB-1065/2020\|EPI_ISL_437065\|2020-0 | ···················· | ···················· | ···················· | ···················· | ···················· | ···················· |
| B | 9333 | hCoV-19/USA/CA-CZB-1066/2020\|EPI_ISL_437066\|2020-0 | ···················· | ···················· | ···················· | ···················· | ···················· | ···················· |
| B | 9334 | hCoV-19/USA/CA-CZB-1067/2020\|EPI_ISL_437067\|2020-0 | ···················· | ···················· | ···················· | ···················· | ···················· | ···················· |
| B | 9335 | hCoV-19/USA/CA-CZB-1068/2020\|EPI_ISL_437068\|2020-0 | ···················· | ···················· | ···················· | ···················· | ···················· | ···················· |
| B | 9336 | hCoV-19/USA/CA-CZB-1071/2020\|EPI_ISL_437071\|2020-0 | ···················· | ···················· | ···················· | ···················· | ···················· | ···················· |
| B | 9337 | hCoV-19/USA/CA-CZB-1072/2020\|EPI_ISL_437072\|2020-0 | ···················· | ···················· | ···················· | ···················· | ···················· | ···················· |
| B | 9338 | hCoV-19/USA/CA-CZB-1073/2020\|EPI_ISL_437073\|2020-0 | ···················· | ···················· | ···················· | ···················· | ···················· | ···················· |
| B | 9339 | hCoV-19/USA/CA-CZB-1074/2020\|EPI_ISL_437074\|2020-0 | ···················· | ···················· | ···················· | ···················· | ···················· | ···················· |
| B | 9340 | hCoV-19/USA/CA-CZB-1075/2020\|EPI_ISL_437075\|2020-0 | ···················· | ···················· | ···················· | ···················· | ···················· | ···················· |
| B | 9341 | hCoV-19/USA/CA-CZB-1076/2020\|EPI_ISL_437076\|2020-0 | ···················· | ···················· | ···················· | ···················· | ···················· | ···················· |
| B | 9342 | hCoV-19/USA/CA-CZB-1077/2020\|EPI_ISL_437077\|2020-0 | ···················· | ···················· | ···················· | ···················· | ···················· | ···················· |
| B | 9343 | hCoV-19/USA/CA-CZB-1079/2020\|EPI_ISL_437079\|2020-0 | ···················· | ···················· | ···················· | ···················· | ···················· | ···················· |
| B | 9344 | hCoV-19/USA/CA-CZB-1081/2020\|EPI_ISL_437081\|2020-0 | ···················· | ···················· | ···················· | ···················· | ···················· | ···················· |
| B | 9345 | hCoV-19/USA/CA-CZB-1082/2020\|EPI_ISL_437082\|2020-0 | ···················· | ···················· | ···················· | ···················· | ···················· | ···················· |
| B | 9346 | hCoV-19/USA/CA-CZB-1083/2020\|EPI_ISL_437083\|2020-0 | ···················· | ···················· | ···················· | ···················· | ···················· | ···················· |
| B | 9347 | hCoV-19/USA/CA-CZB-1084/2020\|EPI_ISL_437084\|2020-0 | ···················· | ···················· | ···················· | ···················· | ···················· | ···················· |
| B | 9348 | hCoV-19/USA/CA-CZB-1085/2020\|EPI_ISL_437085\|2020-0 | ···················· | ···················· | ···················· | ···················· | ···················· | ···················· |
| B | 9349 | hCoV-19/USA/CA-CZB-1087/2020\|EPI_ISL_437087\|2020-0 | ···················· | ···················· | ···················· | ···················· | ···················· | ···················· |
| B | 9350 | hCoV-19/USA/CA-CZB-1088/2020\|EPI_ISL_437088\|2020-0 | ···················· | ···················· | ···················· | ···················· | ···················· | ···················· |
| B | 9351 | hCoV-19/Latvia/010/2020\|EPI_ISL_437089\|2020-03-23 | ···················· | ···················· | ···················· | ···················· | ···················· | ···················· |
| B | 9352 | hCoV-19/England/CAMB-75974/2020\|EPI_ISL_438340\|202 | ···················· | ···················· | ···················· | ···················· | ···················· | ···················· |
| B | 9353 | hCoV-19/England/CAMB-75B14/2020\|EPI_ISL_438342\|202 | ···················· | ···················· | ···················· | ···················· | ···················· | ···················· |
| B | 9354 | hCoV-19/England/CAMB-76287/2020\|EPI_ISL_438343\|202 | ···················· | ···················· | ···················· | ···················· | ···················· | ···················· |
| B | 9355 | hCoV-19/England/CAMB-75E2A/2020\|EPI_ISL_438344\|202 | ···················· | ···················· | ···················· | ···················· | ···················· | ···················· |
| B | 9356 | hCoV-19/England/CAMB-76384/2020\|EPI_ISL_438345\|202 | ···················· | ···················· | ···················· | ···················· | ···················· | ···················· |
| B | 9357 | hCoV-19/England/CAMB-76490/2020\|EPI_ISL_438346\|202 | ···················· | ···················· | ···················· | ···················· | ···················· | ···················· |
| B | 9358 | hCoV-19/England/CAMB-768B2/2020\|EPI_ISL_438347\|202 | ···················· | ···················· | ···················· | ···················· | ···················· | ···················· |
| B | 9359 | hCoV-19/England/CAMB-767D3/2020\|EPI_ISL_438349\|202 | ···················· | ···················· | ···················· | ···················· | ···················· | ···················· |
| B | 9360 | hCoV-19/England/CAMB-768A3/2020\|EPI_ISL_438350\|202 | ···················· | ···················· | ···················· | ···················· | ···················· | ···················· |
| B | 9361 | hCoV-19/England/CAMB-75DA5/2020\|EPI_ISL_438351\|202 | ···················· | ···················· | ···················· | ···················· | ···················· | ···················· |
| B | 9362 | hCoV-19/England/CAMB-75F72/2020\|EPI_ISL_438352\|202 | ···················· | ···················· | ···················· | ···················· | ···················· | ···················· |
| B | 9363 | hCoV-19/England/CAMB-763EE/2020\|EPI_ISL_438354\|202 | ···················· | ···················· | ···················· | ···················· | ···················· | ···················· |
| B | 9364 | hCoV-19/England/CAMB-75D1E/2020\|EPI_ISL_438355\|202 | ···················· | ···················· | ···················· | ···················· | ···················· | ···················· |
| B | 9365 | hCoV-19/England/CAMB-76621/2020\|EPI_ISL_438356\|202 | ···················· | ···················· | ···················· | ···················· | ···················· | ···················· |
| B | 9366 | hCoV-19/England/CAMB-76506/2020\|EPI_ISL_438358\|202 | ···················· | ···················· | ···················· | ···················· | ···················· | ···················· |
| B | 9367 | hCoV-19/England/CAMB-769EC/2020\|EPI_ISL_438359\|202 | ···················· | ·A·················· | ···················· | ···················· | ···················· | ···················· |
| B | 9368 | hCoV-19/England/CAMB-75FCD/2020\|EPI_ISL_438360\|202 | ···················· | ···················· | ···················· | ···················· | ···················· | ···················· |
| B | 9369 | hCoV-19/England/CAMB-7690A/2020\|EPI_ISL_438361\|202 | ···················· | ···················· | ···················· | ···················· | ···················· | ···················· |
| B | 9370 | hCoV-19/England/CAMB-762D2/2020\|EPI_ISL_438362\|202 | ···················· | ···················· | ···················· | ···················· | ···················· | ···················· |
| B | 9371 | hCoV-19/England/CAMB-76560/2020\|EPI_ISL_438363\|202 | ···················· | ···················· | ···················· | ···················· | ···················· | ···················· |
| B | 9372 | hCoV-19/England/CAMB-76463/2020\|EPI_ISL_438365\|202 | ···················· | ···················· | ···················· | ···················· | ···················· | ···················· |
| B | 9373 | hCoV-19/England/CAMB-7622D/2020\|EPI_ISL_438366\|202 | ···················· | ···················· | ···················· | ···················· | ···················· | ···················· |
| B | 9374 | hCoV-19/England/CAMB-75F27/2020\|EPI_ISL_438367\|202 | ···················· | ···················· | ···················· | ···················· | ···················· | ···················· |
| B | 9375 | hCoV-19/England/CAMB-76CD4/2020\|EPI_ISL_438368\|202 | ···················· | ···················· | ···················· | ···················· | ···················· | ···················· |
| B | 9376 | hCoV-19/England/CAMB-762B4/2020\|EPI_ISL_438369\|202 | ···················· | ···················· | ···················· | ···················· | ···················· | ···················· |
| B | 9377 | hCoV-19/England/CAMB-75B50/2020\|EPI_ISL_438370\|202 | ···················· | ···················· | ···················· | ···················· | ···················· | ···················· |
| B | 9378 | hCoV-19/England/CAMB-75BE7/2020\|EPI_ISL_438371\|202 | ···················· | ···················· | ···················· | ···················· | ···················· | ···················· |
| B | 9379 | hCoV-19/England/CAMB-76B40/2020\|EPI_ISL_438372\|202 | ···················· | ···················· | ···················· | ···················· | ···················· | ···················· |
| B | 9380 | hCoV-19/England/CAMB-768EF/2020\|EPI_ISL_438375\|202 | ···················· | ···················· | ···················· | ···················· | ···················· | ···················· |
| B | 9381 | hCoV-19/England/CAMB-76CC5/2020\|EPI_ISL_438376\|202 | ···················· | ···················· | ···················· | ···················· | ···················· | ···················· |
| B | 9382 | hCoV-19/England/CAMB-76A34/2020\|EPI_ISL_438377\|202 | ···················· | ···················· | ···················· | ···················· | ···················· | ···················· |
| B | 9383 | hCoV-19/England/CAMB-75B41/2020\|EPI_ISL_438378\|202 | ···················· | ···················· | ···················· | ···················· | ···················· | ···················· |
| B | 9384 | hCoV-19/England/CAMB-76AAD/2020\|EPI_ISL_438380\|202 | ···················· | ···················· | ···················· | ···················· | ···················· | ···················· |
| B | 9385 | hCoV-19/England/CAMB-76B5F/2020\|EPI_ISL_438381\|202 | ···················· | ···················· | ···················· | ···················· | ···················· | ···················· |
| B | 9386 | hCoV-19/England/CAMB-7675B/2020\|EPI_ISL_438382\|202 | ···················· | ···················· | ···················· | ···················· | ···················· | ···················· |
| B | 9387 | hCoV-19/England/CAMB-767E2/2020\|EPI_ISL_438383\|202 | ···················· | ···················· | ···················· | ···················· | ···················· | ···················· |
| B | 9388 | hCoV-19/England/CAMB-76348/2020\|EPI_ISL_438384\|202 | ···················· | ···················· | ···················· | ···················· | ···················· | ···················· |
| B | 9389 | hCoV-19/England/CAMB-75F45/2020\|EPI_ISL_438385\|202 | ···················· | ···················· | ···················· | ···················· | ···················· | ···················· |
| B | 9390 | hCoV-19/England/CAMB-75E75/2020\|EPI_ISL_438387\|202 | ···················· | ···················· | ···················· | ···················· | ···················· | ···················· |
| B | 9391 | hCoV-19/England/CAMB-75A71/2020\|EPI_ISL_438388\|202 | ···················· | ···················· | ···················· | ···················· | ···················· | ···················· |
| B | 9392 | hCoV-19/England/CAMB-75E0C/2020\|EPI_ISL_438389\|202 | ···················· | ···················· | ···················· | ···················· | ···················· | ···················· |
| B | 9393 | hCoV-19/England/CAMB-76982/2020\|EPI_ISL_438390\|202 | ···················· | ···················· | ···················· | ···················· | ···················· | ···················· |
| B | 9394 | hCoV-19/England/CAMB-71DA9/2020\|EPI_ISL_438391\|202 | ···················· | ···················· | ···················· | ···················· | ···················· | ···················· |
| B | 9395 | hCoV-19/England/CAMB-767A6/2020\|EPI_ISL_438392\|202 | ···················· | ···················· | ···················· | ···················· | ···················· | ···················· |
| B | 9396 | hCoV-19/England/CAMB-7676A/2020\|EPI_ISL_438394\|202 | ···················· | ···················· | ···················· | ···················· | ···················· | ···················· |
| B | 9397 | hCoV-19/England/CAMB-76919/2020\|EPI_ISL_438396\|202 | ···················· | ···················· | ···················· | ···················· | ···················· | ···················· |
| B | 9398 | hCoV-19/England/CAMB-76B31/2020\|EPI_ISL_438398\|202 | ···················· | ···················· | ···················· | ···················· | ···················· | ···················· |
| B | 9399 | hCoV-19/England/CAMB-75F54/2020\|EPI_ISL_438399\|202 | ···················· | ···················· | ···················· | ···················· | ···················· | ···················· |
| B | 9400 | hCoV-19/England/CAMB-7282F/2020\|EPI_ISL_439606\|202 | ···················· | ···················· | ···················· | ···················· | ···················· | ···················· |

| | | | | | | | |
|---|---|---|---|---|---|---|---|
| B | 9401 hCoV-19/England/CAMB-71C60/2020\|EPI_ISL_439607\|202 | ···················· | ···················· | ···················· | ···················· | ···········G········ | ···················· |
| B | 9402 hCoV-19/England/CAMB-7217F/2020\|EPI_ISL_439608\|202 | ···················· | ···················· | ···················· | ···················· | ···················· | ···················· |
| B | 9403 hCoV-19/England/CAMB-726BC/2020\|EPI_ISL_439609\|202 | ···················· | ···················· | ···················· | ···················· | ···················· | ···················· |
| B | 9404 hCoV-19/England/CAMB-71CCA/2020\|EPI_ISL_439610\|202 | ···················· | ···················· | ···················· | ···················· | ···················· | ···················· |
| B | 9405 hCoV-19/England/CAMB-71B45/2020\|EPI_ISL_439611\|202 | ···················· | ···················· | ···················· | ···················· | ···················· | ···················· |
| B | 9406 hCoV-19/England/CAMB-71B72/2020\|EPI_ISL_439612\|202 | ···················· | ···················· | ···················· | ···················· | ···················· | ···················· |
| B | 9407 hCoV-19/England/CAMB-726DA/2020\|EPI_ISL_439613\|202 | ···················· | ···················· | ···················· | ···················· | ···················· | ···················· |
| B | 9408 hCoV-19/England/CAMB-72A83/2020\|EPI_ISL_439614\|202 | ···················· | ···················· | ···················· | ···················· | ···················· | ···················· |
| B | 9409 hCoV-19/England/CAMB-72519/2020\|EPI_ISL_439615\|202 | ···················· | ···················· | ···················· | ···················· | ···················· | ···················· |
| B | 9410 hCoV-19/England/CAMB-722F4/2020\|EPI_ISL_439618\|202 | ···················· | ···················· | ···················· | ···················· | ···················· | ···················· |
| B | 9411 hCoV-19/England/CAMB-71B18/2020\|EPI_ISL_439619\|202 | ···················· | ···················· | ···················· | ···················· | ···················· | ···················· |
| B | 9412 hCoV-19/England/CAMB-71F0D/2020\|EPI_ISL_439627\|202 | ···················· | ···················· | ···················· | ···················· | ···················· | ···················· |
| B | 9413 hCoV-19/Scotland/EDB829/2020\|EPI_ISL_439671\|2020-0 | ···················· | ·A·················· | ···················· | ···················· | ···················· | ···················· |
| B | 9414 hCoV-19/England/LIVE-98F00/2020\|EPI_ISL_439672\|202 | ···················· | ···················· | ···················· | ···················· | ···················· | ···················· |
| B | 9415 hCoV-19/England/LIVE-98F2E/2020\|EPI_ISL_439673\|202 | ···················· | ···················· | ···················· | ···················· | ···················· | ···················· |
| B | 9416 hCoV-19/England/LIVE-98F79/2020\|EPI_ISL_439677\|202 | ···················· | ···················· | ···················· | ···················· | ···················· | ···················· |
| B | 9417 hCoV-19/England/LIVE-98FD3/2020\|EPI_ISL_439681\|202 | ···················· | ···················· | ···················· | ···················· | ···················· | ···················· |
| B | 9418 hCoV-19/England/LIVE-98FF1/2020\|EPI_ISL_439682\|202 | ···················· | ···················· | ···················· | ···················· | ···················· | ···················· |
| B | 9419 hCoV-19/England/LIVE-9900C/2020\|EPI_ISL_439683\|202 | ···················· | ···················· | ···················· | ···················· | ···················· | ···················· |
| B | 9420 hCoV-19/England/LIVE-9901B/2020\|EPI_ISL_439684\|202 | ···················· | ···················· | ···················· | ···················· | ···················· | ···················· |
| B | 9421 hCoV-19/England/LIVE-99048/2020\|EPI_ISL_439687\|202 | ···················· | ···················· | ···················· | ···················· | ···················· | ···················· |
| B | 9422 hCoV-19/England/LIVE-991DC/2020\|EPI_ISL_439697\|202 | ···················· | ···················· | ···················· | ···················· | ···················· | ···················· |
| B | 9423 hCoV-19/England/CAMB-79CA4/2020\|EPI_ISL_440604\|202 | ···················· | ···················· | ···················· | ···················· | ···················· | ···················· |
| B | 9424 hCoV-19/England/CAMB-79F9C/2020\|EPI_ISL_440605\|202 | ···················· | ···················· | ···················· | ···················· | ···················· | ···················· |
| B | 9425 hCoV-19/England/BRIS-1226D8/2020\|EPI_ISL_440653\|20 | ···················· | ···················· | ···················· | ···················· | ···················· | ···················· |
| B | 9426 hCoV-19/England/BRIS-1227E4/2020\|EPI_ISL_440655\|20 | ···················· | ···················· | ···················· | ···················· | ···················· | ···················· |
| B | 9427 hCoV-19/England/BRIS-1227D5/2020\|EPI_ISL_440656\|20 | ···················· | ···················· | ···················· | ···················· | ···················· | ···················· |
| B | 9428 hCoV-19/England/BRIS-12240B/2020\|EPI_ISL_440657\|20 | ···················· | ···················· | ···················· | ···················· | ···················· | ···················· |
| B | 9429 hCoV-19/England/BRIS-122A09/2020\|EPI_ISL_440658\|20 | ···················· | ···················· | ···················· | ···················· | ···················· | ···················· |
| B | 9430 hCoV-19/England/BRIS-12277B/2020\|EPI_ISL_440659\|20 | ···················· | ···················· | ···················· | ···················· | ···················· | ···················· |
| B | 9431 hCoV-19/England/BRIS-122FBF/2020\|EPI_ISL_440660\|20 | ···················· | ···A················ | ···················· | ···················· | ···················· | ···················· |
| B | 9432 hCoV-19/England/BRIS-122F82/2020\|EPI_ISL_440662\|20 | ···················· | ···················· | ···················· | ···················· | ···················· | ···················· |
| B | 9433 hCoV-19/England/BRIS-122ADC/2020\|EPI_ISL_440663\|20 | ···················· | ···················· | ···················· | ···················· | ···················· | ···················· |
| B | 9434 hCoV-19/England/BRIS-122B51/2020\|EPI_ISL_440664\|20 | ···················· | ···················· | ···················· | ···················· | ···················· | ···················· |
| B | 9435 hCoV-19/England/BRIS-122C03/2020\|EPI_ISL_440665\|20 | ···················· | ···················· | ···················· | ···················· | ···················· | ···················· |
| B | 9436 hCoV-19/England/BRIS-122D3D/2020\|EPI_ISL_440666\|20 | ···················· | ···················· | ···················· | ···················· | ···················· | ···················· |
| B | 9437 hCoV-19/England/BRIS-1229C0/2020\|EPI_ISL_440667\|20 | ···················· | ·············A······ | ···················· | ···················· | ···················· | ···················· |
| B | 9438 hCoV-19/England/BRIS-122535/2020\|EPI_ISL_440670\|20 | ···················· | ···················· | ···················· | ···················· | ···················· | ···················· |
| B | 9439 hCoV-19/England/BRIS-122632/2020\|EPI_ISL_440671\|20 | ···················· | ···················· | ···················· | ···················· | ···················· | ···················· |
| B | 9440 hCoV-19/England/BRIS-1224B0/2020\|EPI_ISL_440673\|20 | ···················· | ···················· | ···················· | ···················· | ···················· | ···················· |
| B | 9441 hCoV-19/England/BRIS-122948/2020\|EPI_ISL_440674\|20 | ···················· | ···················· | ···················· | ···················· | ···················· | ···················· |
| B | 9442 hCoV-19/England/BRIS-1223D1/2020\|EPI_ISL_440676\|20 | ···················· | ···················· | ···················· | ···················· | ···················· | ···················· |
| B | 9443 hCoV-19/England/BRIS-12267E/2020\|EPI_ISL_440679\|20 | ···················· | ···················· | ···················· | ···················· | ···················· | ···················· |
| B | 9444 hCoV-19/England/BRIS-122D4C/2020\|EPI_ISL_440681\|20 | ···················· | ···················· | ···················· | ···················· | ···················· | ···················· |
| B | 9445 hCoV-19/England/BRIS-122720/2020\|EPI_ISL_440682\|20 | ···················· | ···················· | ···················· | ···················· | ···················· | ···················· |
| B | 9446 hCoV-19/England/BRIS-122B06/2020\|EPI_ISL_440683\|20 | ········T··········· | ···················· | ···················· | ···················· | ···················· | ···················· |
| B | 9447 hCoV-19/England/BRIS-122A72/2020\|EPI_ISL_440684\|20 | ···················· | ···················· | ···················· | ···················· | ···················· | ···················· |
| B | 9448 hCoV-19/England/BRIS-1226F6/2020\|EPI_ISL_440685\|20 | ···················· | ···················· | ···················· | ···················· | ···················· | ···················· |
| B | 9449 hCoV-19/England/BRIS-12269C/2020\|EPI_ISL_440688\|20 | ···················· | ···················· | ···················· | ···················· | ···················· | ···················· |
| B | 9450 hCoV-19/England/BRIS-1224ED/2020\|EPI_ISL_440689\|20 | ···················· | ···················· | ···················· | ···················· | ···················· | ···················· |
| B | 9451 hCoV-19/England/BRIS-122641/2020\|EPI_ISL_440690\|20 | ···················· | ···················· | ···················· | ···················· | ···················· | ···················· |
| B | 9452 hCoV-19/England/BRIS-1224A1/2020\|EPI_ISL_440691\|20 | ···················· | ···················· | ···················· | ···················· | ···················· | ···················· |
| B | 9453 hCoV-19/England/BRIS-122492/2020\|EPI_ISL_440692\|20 | ···················· | ···················· | ···················· | ···················· | ···················· | ···················· |
| B | 9454 hCoV-19/England/BRIS-122C5E/2020\|EPI_ISL_440694\|20 | ···················· | ···················· | ···················· | ···················· | ···················· | ···················· |
| B | 9455 hCoV-19/England/BRIS-12266F/2020\|EPI_ISL_440695\|20 | ···················· | ···················· | ···················· | ···················· | ···················· | ···················· |
| B | 9456 hCoV-19/England/BRIS-1225AE/2020\|EPI_ISL_440697\|20 | ···················· | ···················· | ···················· | ···················· | ···················· | ···················· |
| B | 9457 hCoV-19/England/BRIS-1227F3/2020\|EPI_ISL_440698\|20 | ···················· | ···················· | ···················· | ···················· | ···················· | ···················· |
| B | 9458 hCoV-19/England/BRIS-122BF7/2020\|EPI_ISL_440699\|20 | ···················· | ···················· | ···················· | ···················· | ···················· | ···················· |
| B | 9459 hCoV-19/England/SHEF-C5F19/2020\|EPI_ISL_441984\|202 | ···················· | ···················· | ···················· | ···················· | ···················· | ···················· |
| B | 9460 hCoV-19/England/SHEF-C6798/2020\|EPI_ISL_441991\|202 | ···················· | ···················· | ···················· | ···················· | ···················· | ···················· |
| B | 9461 hCoV-19/England/SHEF-C6C20/2020\|EPI_ISL_441999\|202 | ···················· | ···················· | ···················· | ···················· | ···················· | ···················· |
| B | 9462 hCoV-19/England/LOND-D52A4/2020\|EPI_ISL_444204\|202 | ···················· | ···················· | ···················· | ···················· | ···················· | ···················· |
| B | 9463 hCoV-19/England/LOND-D52B3/2020\|EPI_ISL_444205\|202 | ···················· | ···················· | ···················· | ···················· | ···················· | ···················· |
| B | 9464 hCoV-19/England/LOND-D52C2/2020\|EPI_ISL_444206\|202 | ···················· | ···················· | ···················· | ···················· | ···················· | ···················· |
| B | 9465 hCoV-19/England/LOND-D52E0/2020\|EPI_ISL_444207\|202 | ···················· | ···················· | ···················· | ···················· | ···················· | ···················· |
| B | 9466 hCoV-19/England/LOND-D52FF/2020\|EPI_ISL_444208\|202 | ···················· | ···················· | ···················· | ···················· | ···················· | ···················· |
| B | 9467 hCoV-19/England/LOND-D530B/2020\|EPI_ISL_444209\|202 | ···················· | ···················· | ···················· | ···················· | ···················· | ···················· |
| B | 9468 hCoV-19/Wales/PHWC-292FC/2020\|EPI_ISL_445507\|2020- | ···················· | ···A················ | ···················· | ···················· | ···················· | ···················· |
| B | 9469 hCoV-19/Wales/PHWC-29335/2020\|EPI_ISL_445510\|2020- | ···················· | ···················· | ···················· | ···················· | ···················· | ···················· |
| B | 9470 hCoV-19/Wales/PHWC-33EA0/2020\|EPI_ISL_446835\|2020- | ···················· | ···A················ | ···················· | ···················· | ···················· | ···················· |
| B | 9471 hCoV-19/Wales/PHWC-33FBC/2020\|EPI_ISL_446851\|2020- | ···················· | ···················· | ···················· | ···················· | ···················· | ···················· |
| B | 9472 hCoV-19/Wales/PHWC-340A9/2020\|EPI_ISL_446866\|2020- | ···················· | ···················· | ···················· | ···················· | ···················· | ···················· |
| B | 9473 hCoV-19/Wales/PHWC-2A167/2020\|EPI_ISL_432184\|2020- | ···················· | ···················· | ···················· | ···················· | ···················· | ···················· |
| B | 9474 hCoV-19/Wales/PHWC-25357/2020\|EPI_ISL_432188\|2020- | ···················· | ···················· | ···················· | ···················· | ···················· | ···················· |
| B | 9475 hCoV-19/Wales/PHWC-29C76/2020\|EPI_ISL_432189\|2020- | ···················· | ···················· | ···················· | ···················· | ···················· | ···················· |
| B | 9476 hCoV-19/Scotland/EDB1433/2020\|EPI_ISL_433446\|2020- | ···················· | ·A·················· | ···················· | ···················· | ···················· | ···················· |
| B | 9477 hCoV-19/Scotland/EDB1436/2020\|EPI_ISL_433447\|2020- | ···················· | ·A·················· | ···················· | ···················· | ···················· | ···················· |
| B | 9478 hCoV-19/Scotland/EDB1437/2020\|EPI_ISL_433448\|2020- | ···················· | ···················· | ···················· | ···················· | ···················· | ···················· |

| | | | | | | | | | |
|---|---|---|---|---|---|---|---|---|---|
| B | 9479 | hCoV-19/Scotland/EDB1444/2020\|EPI_ISL_433449\|2020- | .................... | .A.................. | .................... | .................... | .................... | .................... | |
| B | 9480 | hCoV-19/Scotland/EDB1447/2020\|EPI_ISL_433450\|2020- | .................... | .A.................. | .................... | .................... | .................... | .................... | |
| B | 9481 | hCoV-19/Scotland/EDB1448/2020\|EPI_ISL_433451\|2020- | .................... | .................... | .................... | .................... | .................... | .................... | |
| B | 9482 | hCoV-19/Scotland/EDB1450/2020\|EPI_ISL_433452\|2020- | .................... | .A.................. | .................... | .................... | .................... | .................... | |
| B | 9483 | hCoV-19/Scotland/EDB1452/2020\|EPI_ISL_433453\|2020- | .................... | .A.................. | .................... | .................... | .................... | .................... | |
| B | 9484 | hCoV-19/Scotland/EDB1453/2020\|EPI_ISL_433454\|2020- | .................... | .A.................. | .................... | .................... | .................... | .................... | |
| B | 9485 | hCoV-19/Scotland/EDB1457/2020\|EPI_ISL_433455\|2020- | .................... | .................... | .................... | .................... | .................... | .................... | |
| B | 9486 | hCoV-19/Scotland/EDB1459/2020\|EPI_ISL_433456\|2020- | .................... | .................... | .................... | .................... | .................... | .................... | |
| B | 9487 | hCoV-19/Scotland/EDB1460/2020\|EPI_ISL_433457\|2020- | .................... | .................... | .................... | .................... | .................... | .................... | |
| B | 9488 | hCoV-19/Scotland/EDB1463/2020\|EPI_ISL_433458\|2020- | .................... | .................... | .................... | .................... | .................... | .................... | |
| B | 9489 | hCoV-19/Scotland/EDB1469/2020\|EPI_ISL_433459\|2020- | .................... | .A.................. | .................... | .................... | .................... | .................... | |
| B | 9490 | hCoV-19/Scotland/EDB1470/2020\|EPI_ISL_433460\|2020- | .................... | .................... | .................... | .................... | .................... | .................... | |
| B | 9491 | hCoV-19/Scotland/EDB1471/2020\|EPI_ISL_433461\|2020- | .................... | .A.................. | .................... | .................... | .................... | .................... | |
| B | 9492 | hCoV-19/Scotland/EDB1507/2020\|EPI_ISL_433462\|2020- | .................... | .................... | .................... | .................... | .................... | .................... | |
| B | 9493 | hCoV-19/Scotland/EDB1510/2020\|EPI_ISL_433463\|2020- | .................... | .................... | .................... | .................... | .................... | .................... | |
| B | 9494 | hCoV-19/Scotland/EDB1511/2020\|EPI_ISL_433464\|2020- | .................... | .................... | .................... | .................... | .................... | .................... | |
| B | 9495 | hCoV-19/England/CAMB-7E2E9/2020\|EPI_ISL_433467\|202 | .................... | .................... | .................... | .................... | .................... | .................... | |
| B | 9496 | hCoV-19/England/CAMB-7E304/2020\|EPI_ISL_433469\|202 | .................... | .................... | .................... | .................... | .................... | .................... | |
| B | 9497 | hCoV-19/England/CAMB-7E313/2020\|EPI_ISL_433470\|202 | .................... | .................... | .................... | .................... | .................... | .................... | |
| B | 9498 | hCoV-19/England/CAMB-7E322/2020\|EPI_ISL_433471\|202 | .................... | .................... | .................... | .................... | .................... | .................... | |
| B | 9499 | hCoV-19/England/CAMB-7E331/2020\|EPI_ISL_433472\|202 | .................... | .................... | .................... | .................... | .................... | .................... | |
| B | 9500 | hCoV-19/England/CAMB-7F8E6/2020\|EPI_ISL_433478\|202 | .................... | .................... | .................... | .................... | .................... | .................... | |
| B | 9501 | hCoV-19/England/CAMB-7FB29/2020\|EPI_ISL_433479\|202 | .................... | .................... | .................... | .................... | .................... | .................... | |
| B | 9502 | hCoV-19/England/CAMB-7FB47/2020\|EPI_ISL_433480\|202 | .................... | .................... | .................... | .................... | .................... | .................... | |
| B | 9503 | hCoV-19/England/CAMB-7FB56/2020\|EPI_ISL_433481\|202 | .................... | .................... | .................... | .................... | .................... | .................... | |
| B | 9504 | hCoV-19/England/CAMB-7FB65/2020\|EPI_ISL_433482\|202 | .................... | .................... | .................... | .................... | .................... | .................... | |
| B | 9505 | hCoV-19/England/CAMB-7FB74/2020\|EPI_ISL_433483\|202 | .................... | .................... | .................... | .................... | .................... | .................... | |
| B | 9506 | hCoV-19/England/CAMB-7FB83/2020\|EPI_ISL_433484\|202 | .................... | .................... | .................... | .................... | .................... | .................... | |
| B | 9507 | hCoV-19/England/CAMB-7FB92/2020\|EPI_ISL_433485\|202 | .................... | .................... | .................... | .................... | .................... | .................... | |
| B | 9508 | hCoV-19/England/CAMB-7FBA1/2020\|EPI_ISL_433486\|202 | .................... | .................... | .................... | .................... | .................... | .................... | |
| B | 9509 | hCoV-19/England/CAMB-7FBB0/2020\|EPI_ISL_433487\|202 | .................... | .................... | .................... | .................... | .................... | .................... | |
| B | 9510 | hCoV-19/England/CAMB-7FBCF/2020\|EPI_ISL_433488\|202 | .................... | .................... | .................... | .................... | .................... | .................... | |
| B | 9511 | hCoV-19/England/CAMB-7FBDE/2020\|EPI_ISL_433489\|202 | .................... | .................... | .................... | .................... | .................... | .................... | |
| B | 9513 | hCoV-19/USA/TX-HMH071/2020\|EPI_ISL_434780\|2020-03- | .................... | .................... | .................... | .................... | ..............N..... | ..N................. | |
| B | 9515 | hCoV-19/USA/TX-HMH074/2020\|EPI_ISL_434783\|2020-03- | .................... | .................... | .................... | .................... | .................... | .................... | |
| B | 9517 | hCoV-19/USA/TX-HMH076/2020\|EPI_ISL_434785\|2020-03- | .................... | .................... | .................... | .................... | .................... | .................... | |
| B | 9518 | hCoV-19/USA/TX-HMH078/2020\|EPI_ISL_434787\|2020-03- | .................... | .................... | .................... | .................... | .................... | .................... | |
| B | 9519 | hCoV-19/USA/TX-HMH079/2020\|EPI_ISL_434788\|2020-03- | .................... | .................... | .................... | .................... | .................... | .................... | |
| B | 9525 | hCoV-19/USA/TX-HMH093/2020\|EPI_ISL_434798\|2020-03- | .................... | .................... | .................... | .................... | ..............N..... | NN.N................ | 1 |
| B | 9527 | hCoV-19/Denmark/ALAB-SSI-619/2020\|EPI_ISL_437000\|2 | .................... | .................... | .................... | .................... | .................... | .................... | |
| B | 9528 | hCoV-19/Denmark/ALAB-SSI-620/2020\|EPI_ISL_437001\|2 | .................... | .................... | .................... | .................... | .................... | .................... | |
| B | 9529 | hCoV-19/Denmark/ALAB-SSI-623/2020\|EPI_ISL_437003\|2 | .................... | .................... | .................... | .................... | .................... | .................... | |
| B | 9530 | hCoV-19/Denmark/ALAB-SSI-624/2020\|EPI_ISL_437004\|2 | .................... | .................... | .................... | .................... | .................... | .................... | |
| B | 9531 | hCoV-19/Denmark/ALAB-SSI-626/2020\|EPI_ISL_437006\|2 | .................... | .................... | .................... | .................... | .................... | .................... | |
| B | 9532 | hCoV-19/Denmark/ALAB-SSI-628/2020\|EPI_ISL_437007\|2 | .................... | .................... | .................... | .................... | .................... | .................... | |
| B | 9533 | hCoV-19/England/CAMB-71D9A/2020\|EPI_ISL_438300\|202 | .................... | .................... | .................... | .................... | .................... | .................... | |
| B | 9534 | hCoV-19/England/CAMB-75C7B/2020\|EPI_ISL_438301\|202 | .................... | .................... | .................... | .................... | .................... | .................... | |
| B | 9535 | hCoV-19/England/CAMB-761A8/2020\|EPI_ISL_438302\|202 | .................... | .................... | .................... | .................... | .................... | .................... | |
| B | 9536 | hCoV-19/England/CAMB-769FB/2020\|EPI_ISL_438303\|202 | .................... | .................... | .................... | .................... | .................... | .................... | |
| B | 9537 | hCoV-19/England/CAMB-75B23/2020\|EPI_ISL_438305\|202 | .................... | .................... | .................... | .................... | .................... | .................... | |
| B | 9538 | hCoV-19/England/CAMB-76C89/2020\|EPI_ISL_438306\|202 | .................... | .................... | .................... | .................... | .................... | .................... | |
| B | 9539 | hCoV-19/England/CAMB-75BAB/2020\|EPI_ISL_438307\|202 | .................... | .................... | .................... | .................... | .................... | .................... | |
| B | 9540 | hCoV-19/England/CAMB-7632A/2020\|EPI_ISL_438308\|202 | .................... | .................... | .................... | .................... | .................... | .................... | |
| B | 9541 | hCoV-19/England/CAMB-76ABC/2020\|EPI_ISL_438309\|202 | .................... | .............A...... | .................... | .................... | .................... | .................... | |
| B | 9542 | hCoV-19/England/CAMB-763C0/2020\|EPI_ISL_438310\|202 | .................... | .................... | .................... | .................... | .................... | .................... | |
| B | 9543 | hCoV-19/England/CAMB-75ABD/2020\|EPI_ISL_438311\|202 | .................... | .................... | .................... | .................... | .................... | .................... | |
| B | 9544 | hCoV-19/England/CAMB-75E93/2020\|EPI_ISL_438312\|202 | .................... | .................... | .................... | .................... | .................... | .................... | |
| B | 9545 | hCoV-19/England/CAMB-75C4E/2020\|EPI_ISL_438315\|202 | .................... | .................... | .................... | .................... | .................... | .................... | |
| B | 9546 | hCoV-19/England/CAMB-7618A/2020\|EPI_ISL_438316\|202 | .................... | .................... | .................... | .................... | .................... | .................... | |
| B | 9547 | hCoV-19/England/CAMB-76A8F/2020\|EPI_ISL_438318\|202 | .................... | .................... | .................... | .................... | .................... | .................... | |
| B | 9548 | hCoV-19/England/CAMB-75AF9/2020\|EPI_ISL_438319\|202 | .................... | .................... | .................... | .................... | .................... | .................... | |
| B | 9549 | hCoV-19/England/CAMB-75E39/2020\|EPI_ISL_438321\|202 | .................... | .................... | .................... | .................... | .................... | .................... | |
| B | 9550 | hCoV-19/England/CAMB-76955/2020\|EPI_ISL_438322\|202 | .................... | ...................A | .................... | .................... | .................... | .................... | 1 |
| B | 9551 | hCoV-19/England/CAMB-76551/2020\|EPI_ISL_438323\|202 | .................... | .................... | .................... | .................... | .................... | .................... | |
| B | 9552 | hCoV-19/England/CAMB-75D5A/2020\|EPI_ISL_438324\|202 | .................... | .................... | .................... | .................... | .................... | .................... | |
| B | 9553 | hCoV-19/England/CAMB-768D0/2020\|EPI_ISL_438325\|202 | .................... | .................... | .................... | .................... | .................... | .................... | |
| B | 9554 | hCoV-19/England/CAMB-769BF/2020\|EPI_ISL_438326\|202 | .................... | .................... | .................... | .................... | .................... | .................... | |
| B | 9555 | hCoV-19/England/CAMB-71DD6/2020\|EPI_ISL_438327\|202 | .................... | .................... | .................... | .................... | .................... | .................... | |
| B | 9556 | hCoV-19/England/CAMB-76A61/2020\|EPI_ISL_438329\|202 | .................... | .................... | .................... | .................... | .................... | .................... | |
| B | 9557 | hCoV-19/England/CAMB-75BF6/2020\|EPI_ISL_438330\|202 | .................... | .................... | .................... | .................... | .................... | .................... | |
| B | 9558 | hCoV-19/England/CAMB-762F0/2020\|EPI_ISL_438332\|202 | .................... | .................... | .................... | .................... | .................... | .................... | |
| B | 9559 | hCoV-19/England/CAMB-76269/2020\|EPI_ISL_438333\|202 | .................... | .................... | .................... | .................... | .................... | .................... | |
| B | 9560 | hCoV-19/England/CAMB-75D96/2020\|EPI_ISL_438334\|202 | .................... | .................... | .................... | .................... | .................... | .................... | |
| B | 9561 | hCoV-19/England/CAMB-7672E/2020\|EPI_ISL_438337\|202 | .................... | .................... | .................... | .................... | .................... | .................... | |
| B | 9562 | hCoV-19/England/CAMB-75BBA/2020\|EPI_ISL_438338\|202 | .................... | .................... | .................... | .................... | .................... | .................... | |
| B | 9563 | hCoV-19/England/CAMB-71BFA/2020\|EPI_ISL_439601\|202 | .................... | .................... | .................... | .................... | .................... | .................... | |
| B | 9564 | hCoV-19/England/CAMB-71E4C/2020\|EPI_ISL_439602\|202 | .................... | .................... | .................... | .................... | .................... | .................... | |
| B | 9565 | hCoV-19/England/CAMB-726CB/2020\|EPI_ISL_439604\|202 | .................... | .................... | .................... | .................... | .................... | .................... | |

| | | | | | | | | |
|---|---|---|---|---|---|---|---|---|
| B | 9566 | hCoV-19/England/CAMB-71B27/2020\|EPI_ISL_439620\|202 | ···················· | ···················· | ···················· | ···················· | ···················· | ···················· |
| B | 9567 | hCoV-19/England/CAMB-72467/2020\|EPI_ISL_439621\|202 | ···················· | ···················· | ···················· | ···················· | ···················· | ···················· |
| B | 9568 | hCoV-19/England/CAMB-72221/2020\|EPI_ISL_439622\|202 | ···················· | ···················· | ···················· | ···················· | ···················· | ···················· |
| B | 9569 | hCoV-19/England/CAMB-71CF7/2020\|EPI_ISL_439623\|202 | ···················· | ···················· | ···················· | ···················· | ···················· | ···················· |
| B | 9570 | hCoV-19/England/CAMB-7219D/2020\|EPI_ISL_439625\|202 | ···················· | ···················· | ···················· | ···················· | ···················· | ···················· |
| B | 9571 | hCoV-19/England/CAMB-71F49/2020\|EPI_ISL_439626\|202 | ···················· | ···················· | ···················· | ···················· | ···················· | ···················· |
| B | 9572 | hCoV-19/England/CAMB-725FB/2020\|EPI_ISL_439628\|202 | ···················· | ···················· | ···················· | ···················· | ···················· | ···················· |
| B | 9573 | hCoV-19/England/CAMB-7287A/2020\|EPI_ISL_439630\|202 | ···················· | ···················· | ···················· | ···················· | ···················· | ···················· |
| B | 9574 | hCoV-19/England/CAMB-72045/2020\|EPI_ISL_439631\|202 | ···················· | ···················· | ···················· | ···················· | ···················· | ···················· |
| B | 9575 | hCoV-19/England/CAMB-71E79/2020\|EPI_ISL_439633\|202 | ···················· | ···················· | ···················· | ···················· | ···················· | ···················· |
| B | 9576 | hCoV-19/England/CAMB-72537/2020\|EPI_ISL_439634\|202 | ···················· | ···················· | ···················· | ···················· | ···················· | ···················· |
| B | 9577 | hCoV-19/England/CAMB-72731/2020\|EPI_ISL_439635\|202 | ···················· | ···················· | ···················· | ···················· | ···················· | ···················· |
| B | 9578 | hCoV-19/England/CAMB-7242B/2020\|EPI_ISL_439636\|202 | ···················· | ···················· | ···················· | ···················· | ···················· | ···················· |
| B | 9579 | hCoV-19/England/CAMB-71C9D/2020\|EPI_ISL_439637\|202 | ···················· | ···················· | ···················· | ···················· | ···················· | ···················· |
| B | 9580 | hCoV-19/England/CAMB-7277D/2020\|EPI_ISL_439638\|202 | ···················· | ···················· | ···················· | ···················· | ···················· | ···················· |
| B | 9581 | hCoV-19/England/CAMB-728A7/2020\|EPI_ISL_439639\|202 | ···················· | ···················· | ···················· | ···················· | ···················· | ···················· |
| B | 9582 | hCoV-19/England/CAMB-72133/2020\|EPI_ISL_439640\|202 | ···················· | ···················· | ···················· | ···················· | ···················· | ···················· |
| B | 9583 | hCoV-19/England/CAMB-72A47/2020\|EPI_ISL_439641\|202 | ···················· | ···················· | ···················· | ···················· | ···················· | ···················· |
| B | 9584 | hCoV-19/England/CAMB-7241C/2020\|EPI_ISL_439642\|202 | ···················· | ···················· | ···················· | ···················· | ···················· | ···················· |
| B | 9585 | hCoV-19/England/CAMB-72634/2020\|EPI_ISL_439643\|202 | ···················· | ···················· | ···················· | ···················· | ···················· | ···················· |
| B | 9586 | hCoV-19/England/CAMB-72740/2020\|EPI_ISL_439644\|202 | ···················· | ···················· | ···················· | ···················· | ···················· | ···················· |
| B | 9587 | hCoV-19/England/CAMB-72722/2020\|EPI_ISL_439645\|202 | ···················· | ···················· | ···················· | ···················· | ···················· | ···················· |
| B | 9588 | hCoV-19/England/CAMB-71D21/2020\|EPI_ISL_439646\|202 | ···················· | ···················· | ···················· | ···················· | ···················· | ···················· |
| B | 9589 | hCoV-19/England/CAMB-71A93/2020\|EPI_ISL_439647\|202 | ···················· | ···················· | ···················· | ···················· | ···················· | ···················· |
| B | 9590 | hCoV-19/England/CAMB-72625/2020\|EPI_ISL_439648\|202 | ···················· | ···················· | ···················· | ···················· | ···················· | ···················· |
| B | 9591 | hCoV-19/England/CAMB-71EC4/2020\|EPI_ISL_439649\|202 | ···················· | ···················· | ···················· | ···················· | ···················· | ···················· |
| B | 9592 | hCoV-19/England/CAMB-71E3D/2020\|EPI_ISL_439652\|202 | ···················· | ············A······· | ···················· | ···················· | ···················· | ···················· |
| B | 9593 | hCoV-19/England/CAMB-721D9/2020\|EPI_ISL_439653\|202 | ···················· | ···················· | ···················· | ···················· | ···················· | ···················· |
| B | 9594 | hCoV-19/England/CAMB-721CA/2020\|EPI_ISL_439655\|202 | ···················· | ···················· | ···················· | ···················· | ···················· | ···················· |
| B | 9595 | hCoV-19/England/CAMB-71F85/2020\|EPI_ISL_439656\|202 | ···················· | ···················· | ···················· | ···················· | ···················· | ···················· |
| B | 9596 | hCoV-19/England/CAMB-721F7/2020\|EPI_ISL_439657\|202 | ···················· | ···················· | ···················· | ···················· | ···················· | ···················· |
| B | 9597 | hCoV-19/England/CAMB-71AC0/2020\|EPI_ISL_439659\|202 | ···················· | ···················· | ···················· | ···················· | ···················· | ···················· |
| B | 9598 | hCoV-19/England/CAMB-72142/2020\|EPI_ISL_439660\|202 | ···················· | ···················· | ···················· | ···················· | ···················· | ···················· |
| B | 9599 | hCoV-19/England/CAMB-72081/2020\|EPI_ISL_439662\|202 | ···················· | ···················· | ···················· | ···················· | ···················· | ···················· |
| B | 9600 | hCoV-19/England/CAMB-71B54/2020\|EPI_ISL_439664\|202 | ···················· | ···················· | ···················· | ···················· | ···················· | ···················· |
| B | 9601 | hCoV-19/Scotland/EDB3925/2020\|EPI_ISL_439665\|2020- | ···················· | ·A·················· | ···················· | ···················· | ···················· | ···················· |
| B | 9602 | hCoV-19/Scotland/EDB3941/2020\|EPI_ISL_439666\|2020- | ···················· | ·A·················· | ···················· | ···················· | ···················· | ···················· |
| B | 9603 | hCoV-19/Scotland/EDB3943/2020\|EPI_ISL_439668\|2020- | ···················· | ···················· | ···················· | ···················· | ···················· | ···················· |
| B | 9605 | hCoV-19/England/CAMB-79FF6/2020\|EPI_ISL_440600\|202 | ···················· | ···················· | ···················· | ···················· | ···················· | ···················· |
| B | 9606 | hCoV-19/England/CAMB-77D2B/2020\|EPI_ISL_440601\|202 | ···················· | ···················· | ···················· | ···················· | ···················· | ···················· |
| B | 9607 | hCoV-19/England/CAMB-79EF9/2020\|EPI_ISL_440603\|202 | ···················· | ···················· | ···················· | ···················· | ···················· | ···················· |
| B | 9608 | hCoV-19/England/CAMB-79E62/2020\|EPI_ISL_440606\|202 | ···················· | ···················· | ···················· | ···················· | ···················· | ···················· |
| B | 9609 | hCoV-19/England/CAMB-7A380/2020\|EPI_ISL_440607\|202 | ···················· | ···················· | ···················· | ···················· | ···················· | ···················· |
| B | 9610 | hCoV-19/England/CAMB-79020/2020\|EPI_ISL_440608\|202 | ···················· | ···················· | ···················· | ···················· | ···················· | ···················· |
| B | 9611 | hCoV-19/England/CAMB-7A098/2020\|EPI_ISL_440610\|202 | ···················· | ···················· | ···················· | ···················· | ···················· | ···················· |
| B | 9612 | hCoV-19/England/CAMB-7A362/2020\|EPI_ISL_440611\|202 | ···················· | ···················· | ···················· | ···················· | ···················· | ···················· |
| B | 9613 | hCoV-19/England/CAMB-7A0C5/2020\|EPI_ISL_440612\|202 | ···················· | ···················· | ···················· | ···················· | ···················· | ···················· |
| B | 9614 | hCoV-19/England/CAMB-79EBD/2020\|EPI_ISL_440613\|202 | ···················· | ···················· | ···················· | ···················· | ···················· | ···················· |
| B | 9615 | hCoV-19/England/CAMB-79D38/2020\|EPI_ISL_440615\|202 | ···················· | ···················· | ···················· | ···················· | ···················· | ···················· |
| B | 9616 | hCoV-19/England/CAMB-7B5C5/2020\|EPI_ISL_440616\|202 | ···················· | ···················· | ···················· | ···················· | ···················· | ···················· |
| B | 9617 | hCoV-19/England/CAMB-79E9F/2020\|EPI_ISL_440617\|202 | ···················· | ···················· | ···················· | ···················· | ···················· | ···················· |
| B | 9618 | hCoV-19/England/CAMB-7A64B/2020\|EPI_ISL_440618\|202 | ···················· | ···················· | ···················· | ···················· | ······T············· | ···················· |
| B | 9619 | hCoV-19/England/CAMB-79E26/2020\|EPI_ISL_440619\|202 | ···················· | ···················· | ···················· | ···················· | ···················· | ···················· |
| B | 9620 | hCoV-19/England/CAMB-79D83/2020\|EPI_ISL_440620\|202 | ···················· | ···················· | ···················· | ···················· | ···················· | ···················· |
| B | 9621 | hCoV-19/England/CAMB-79B89/2020\|EPI_ISL_440621\|202 | ···················· | ···················· | ···················· | ···················· | ···················· | ···················· |
| B | 9622 | hCoV-19/England/CAMB-78F24/2020\|EPI_ISL_440622\|202 | ···················· | ···················· | ···················· | ···················· | ···················· | ···················· |
| B | 9623 | hCoV-19/England/BRIS-122966/2020\|EPI_ISL_440624\|20 | ···················· | ···················· | ···················· | ···················· | ···················· | ···················· |
| B | 9624 | hCoV-19/England/BRIS-122E0D/2020\|EPI_ISL_440626\|20 | ···················· | ···················· | ···················· | ···················· | ···················· | ···················· |
| B | 9625 | hCoV-19/England/BRIS-12290C/2020\|EPI_ISL_440628\|20 | ···················· | ···················· | ···················· | ···················· | ···················· | ···················· |
| B | 9626 | hCoV-19/England/BRIS-122B24/2020\|EPI_ISL_440632\|20 | ···················· | ···················· | ···················· | ···················· | ···················· | ···················· |
| B | 9627 | hCoV-19/England/BRIS-122F0A/2020\|EPI_ISL_440634\|20 | ···················· | ···················· | ···················· | ···················· | ···················· | ···················· |
| B | 9628 | hCoV-19/England/BRIS-122ED0/2020\|EPI_ISL_440635\|20 | ···················· | ···················· | ···················· | ···················· | ···················· | ···················· |
| B | 9629 | hCoV-19/England/BRIS-12283C/2020\|EPI_ISL_440636\|20 | ···················· | ···················· | ···················· | ···················· | ···················· | ···················· |
| B | 9630 | hCoV-19/England/BRIS-122F37/2020\|EPI_ISL_440639\|20 | ···················· | ···················· | ···················· | ···················· | ···················· | ···················· |
| B | 9631 | hCoV-19/England/BRIS-122C6D/2020\|EPI_ISL_440640\|20 | ···················· | ···················· | ···················· | ···················· | ···················· | ···················· |
| B | 9632 | hCoV-19/England/BRIS-122EB2/2020\|EPI_ISL_440641\|20 | ···················· | ···················· | ···················· | ···················· | ···················· | ···················· |
| B | 9633 | hCoV-19/England/BRIS-122580/2020\|EPI_ISL_440642\|20 | ···················· | ···················· | ···················· | ···················· | ···················· | ···················· |
| B | 9634 | hCoV-19/England/BRIS-122553/2020\|EPI_ISL_440643\|20 | ···················· | ···················· | ···················· | ···················· | ···················· | ···················· |
| B | 9635 | hCoV-19/England/BRIS-1223C2/2020\|EPI_ISL_440645\|20 | ···················· | ···················· | ···················· | ···················· | ···················· | ···················· |
| B | 9636 | hCoV-19/England/BRIS-122B15/2020\|EPI_ISL_440646\|20 | ···················· | ···················· | ···················· | ···················· | ···················· | ···················· |
| B | 9637 | hCoV-19/England/BRIS-122FDD/2020\|EPI_ISL_440647\|20 | ···················· | ···················· | ···················· | ···················· | ···················· | ···················· |
| B | 9638 | hCoV-19/England/BRIS-122DB5/2020\|EPI_ISL_440649\|20 | ···················· | ···················· | ···················· | ···················· | ···················· | ···················· |
| B | 9639 | hCoV-19/England/BRIS-122AAF/2020\|EPI_ISL_440650\|20 | ···················· | ···················· | ···················· | ···················· | ···················· | ···················· |
| B | 9640 | hCoV-19/England/SHEF-C18F4/2020\|EPI_ISL_441913\|202 | ···················· | ···················· | ···················· | ···················· | ···················· | ···················· |
| B | 9641 | hCoV-19/England/SHEF-C1C8F/2020\|EPI_ISL_441916\|202 | ···················· | ···················· | ···················· | ···················· | ···················· | ···················· |
| B | 9642 | hCoV-19/England/SHEF-C291E/2020\|EPI_ISL_441932\|202 | ···················· | ···················· | ···················· | ···················· | ···················· | ···················· |
| B | 9643 | hCoV-19/England/SHEF-C2D8B/2020\|EPI_ISL_441935\|202 | ···················· | ···················· | ···················· | ···················· | ···················· | ···················· |
| B | 9644 | hCoV-19/England/SHEF-C342B/2020\|EPI_ISL_441942\|202 | ···················· | ···················· | ···················· | ···················· | ···················· | ···················· |

| | | | | | | | | |
|---|---|---|---|---|---|---|---|---|
| B | 9645 | hCoV-19/England/SHEF-C3B62/2020\|EPI_ISL_441952\|202 | ···················· | ···················· | ···················· | ···················· | ···················· | ···················· |
| B | 9646 | hCoV-19/England/SHEF-C3E78/2020\|EPI_ISL_441955\|202 | ···················· | ···················· | ···················· | ···················· | ···················· | ···················· |
| B | 9647 | hCoV-19/England/SHEF-C5650/2020\|EPI_ISL_441975\|202 | ···················· | ···················· | ···················· | ···················· | ···················· | ···················· |
| B | 9648 | hCoV-19/England/SHEF-C5A81/2020\|EPI_ISL_441981\|202 | ···················· | ···················· | ···················· | ···················· | ···················· | ···················· |
| B | 9649 | hCoV-19/England/LOND-D41E4/2020\|EPI_ISL_444100\|202 | ···················· | ···················· | ···················· | ···················· | ···················· | ···················· |
| B | 9650 | hCoV-19/England/LOND-D41F3/2020\|EPI_ISL_444101\|202 | ···················· | ···················· | ···················· | ···················· | ···················· | ···················· |
| B | 9651 | hCoV-19/England/LOND-D420F/2020\|EPI_ISL_444102\|202 | ···················· | ···················· | ···················· | ···················· | ···················· | ···················· |
| B | 9652 | hCoV-19/England/LOND-D421E/2020\|EPI_ISL_444103\|202 | ···················· | ···················· | ···················· | ···················· | ···················· | ···················· |
| B | 9653 | hCoV-19/England/LOND-D422D/2020\|EPI_ISL_444104\|202 | ···················· | ···················· | ···················· | ···················· | ···················· | ···················· |
| B | 9654 | hCoV-19/England/LOND-D423C/2020\|EPI_ISL_444105\|202 | ···················· | ···················· | ···················· | ···················· | ···················· | ···················· |
| B | 9655 | hCoV-19/England/LOND-D424B/2020\|EPI_ISL_444106\|202 | ···················· | ···················· | ···················· | ···················· | ···················· | ···················· |
| B | 9656 | hCoV-19/England/LOND-D4287/2020\|EPI_ISL_444108\|202 | ···················· | ···················· | ···················· | ···················· | ···················· | ···················· |
| B | 9657 | hCoV-19/England/LOND-D4296/2020\|EPI_ISL_444109\|202 | ···················· | ···················· | ···················· | ···················· | ···················· | ···················· |
| B | 9658 | hCoV-19/England/LOND-D42A5/2020\|EPI_ISL_444110\|202 | ···················· | ···················· | ···················· | ···················· | ···················· | ···················· |
| B | 9659 | hCoV-19/England/LOND-D42B4/2020\|EPI_ISL_444111\|202 | ···················· | ···················· | ···················· | ···················· | ···················· | ···················· |
| B | 9660 | hCoV-19/England/LOND-D42C3/2020\|EPI_ISL_444112\|202 | ···················· | ···················· | ···················· | ···················· | ···················· | ···················· |
| B | 9661 | hCoV-19/England/LOND-D42E1/2020\|EPI_ISL_444114\|202 | ···················· | ···················· | ···················· | ···················· | ···················· | ···················· |
| B | 9662 | hCoV-19/England/LOND-D431B/2020\|EPI_ISL_444116\|202 | ···················· | ···················· | ···················· | ···················· | ···················· | ···················· |
| B | 9663 | hCoV-19/England/LOND-D432A/2020\|EPI_ISL_444117\|202 | ···················· | ···················· | ···················· | ···················· | ···················· | ···················· |
| B | 9664 | hCoV-19/England/LOND-D4366/2020\|EPI_ISL_444120\|202 | ···················· | ···················· | ···················· | ···················· | ···················· | ···················· |
| B | 9665 | hCoV-19/England/LOND-D4445/2020\|EPI_ISL_444122\|202 | ···················· | ···················· | ···················· | ···················· | ···················· | ···················· |
| B | 9666 | hCoV-19/England/LOND-D4454/2020\|EPI_ISL_444123\|202 | ···················· | ···················· | ···················· | ···················· | ···················· | ···················· |
| B | 9667 | hCoV-19/England/LOND-D4472/2020\|EPI_ISL_444124\|202 | ···················· | ···················· | ···················· | ···················· | ···················· | ···················· |
| B | 9668 | hCoV-19/England/LOND-D4481/2020\|EPI_ISL_444125\|202 | ···················· | ···················· | ···················· | ···················· | ···················· | ···················· |
| B | 9669 | hCoV-19/England/LOND-D4490/2020\|EPI_ISL_444126\|202 | ···················· | ···················· | ···················· | ···················· | ···················· | ···················· |
| B | 9670 | hCoV-19/England/LOND-D44AF/2020\|EPI_ISL_444127\|202 | ···················· | ···················· | ···················· | ···················· | ···················· | ···················· |
| B | 9671 | hCoV-19/England/LOND-D44BE/2020\|EPI_ISL_444128\|202 | ···················· | ···················· | ···················· | ···················· | ···················· | ···················· |
| B | 9672 | hCoV-19/England/LOND-D44CD/2020\|EPI_ISL_444129\|202 | ···················· | ···················· | ···················· | ···················· | ···················· | ···················· |
| B | 9673 | hCoV-19/England/LOND-D44DC/2020\|EPI_ISL_444130\|202 | ···················· | ···················· | ···················· | ···················· | ···················· | ···················· |
| B | 9674 | hCoV-19/England/LOND-D4560/2020\|EPI_ISL_444133\|202 | ···················· | ···················· | ···················· | ···················· | ···················· | ···················· |
| B | 9675 | hCoV-19/England/LOND-D457F/2020\|EPI_ISL_444134\|202 | ···················· | ···················· | ···················· | ···················· | ···················· | ···················· |
| B | 9676 | hCoV-19/England/LOND-D45AC/2020\|EPI_ISL_444137\|202 | ···················· | ···················· | ···················· | ···················· | ··············N····· | NN·················· |
| B | 9677 | hCoV-19/England/LOND-D4982/2020\|EPI_ISL_444160\|202 | ···················· | ···················· | ···················· | ···················· | ···················· | ···················· |
| B | 9678 | hCoV-19/England/LOND-D4991/2020\|EPI_ISL_444161\|202 | ···················· | ···················· | ···················· | ···················· | ···················· | ···················· |
| B | 9679 | hCoV-19/England/LOND-D49A0/2020\|EPI_ISL_444162\|202 | ···················· | ···················· | ···················· | ···················· | ···················· | ···················· |
| B | 9680 | hCoV-19/England/LOND-D49BF/2020\|EPI_ISL_444163\|202 | ···················· | ···················· | ···················· | ···················· | ···················· | ···················· |
| B | 9681 | hCoV-19/England/LOND-D49CE/2020\|EPI_ISL_444164\|202 | ···················· | ···················· | ···················· | ···················· | ···················· | ···················· |
| B | 9682 | hCoV-19/England/LOND-D49DD/2020\|EPI_ISL_444165\|202 | ···················· | ···················· | ···················· | ···················· | ···················· | ···················· |
| B | 9683 | hCoV-19/England/LOND-D49EC/2020\|EPI_ISL_444166\|202 | ···················· | ···················· | ···················· | ···················· | ···················· | ···················· |
| B | 9684 | hCoV-19/England/LOND-D49FB/2020\|EPI_ISL_444167\|202 | ···················· | ···················· | ···················· | ···················· | ···················· | ···················· |
| B | 9685 | hCoV-19/England/LOND-D4A07/2020\|EPI_ISL_444168\|202 | ···················· | ···················· | ···················· | ···················· | ···················· | ···················· |
| B | 9686 | hCoV-19/England/LOND-D4A16/2020\|EPI_ISL_444169\|202 | ···················· | ···················· | ···················· | ···················· | ···················· | ···················· |
| B | 9687 | hCoV-19/England/LOND-D4B40/2020\|EPI_ISL_444180\|202 | ···················· | ···················· | ···················· | ···················· | ···················· | ···················· |
| B | 9688 | hCoV-19/England/LOND-D4B5F/2020\|EPI_ISL_444181\|202 | ···················· | ···················· | ···················· | ···················· | ···················· | ···················· |
| B | 9689 | hCoV-19/England/LOND-D4B8C/2020\|EPI_ISL_444182\|202 | ···················· | ···················· | ···················· | ···················· | ···················· | ···················· |
| B | 9690 | hCoV-19/England/LOND-D4D68/2020\|EPI_ISL_444185\|202 | ···················· | ···················· | ···················· | ···················· | ···················· | ···················· |
| B | 9691 | hCoV-19/England/LOND-D4FAE/2020\|EPI_ISL_444188\|202 | ···················· | ···················· | ···················· | ···················· | ···················· | ···················· |
| B | 9692 | hCoV-19/England/LOND-D4FDB/2020\|EPI_ISL_444189\|202 | ···················· | ···················· | ···················· | ···················· | ···················· | ···················· |
| B | 9693 | hCoV-19/England/LOND-D4FEA/2020\|EPI_ISL_444190\|202 | ···················· | ···················· | ···················· | ···················· | ···················· | ···················· |
| B | 9694 | hCoV-19/England/LOND-D4FF9/2020\|EPI_ISL_444191\|202 | ···················· | ···················· | ···················· | ···················· | ···················· | ···················· |
| B | 9695 | hCoV-19/England/LOND-D5004/2020\|EPI_ISL_444192\|202 | ···················· | ···················· | ···················· | ···················· | ···················· | ···················· |
| B | 9696 | hCoV-19/England/LOND-D5013/2020\|EPI_ISL_444193\|202 | ···················· | ···················· | ···················· | ···················· | ···················· | ···················· |
| B | 9697 | hCoV-19/England/LOND-D5022/2020\|EPI_ISL_444194\|202 | ···················· | ···················· | ···················· | ···················· | ···················· | ···················· |
| B | 9698 | hCoV-19/England/LOND-D5031/2020\|EPI_ISL_444195\|202 | ···················· | ···················· | ···················· | ···················· | ···················· | ···················· |
| B | 9699 | hCoV-19/England/LOND-D5040/2020\|EPI_ISL_444196\|202 | ···················· | ···················· | ···················· | ···················· | ···················· | ···················· |
| B | 9700 | hCoV-19/England/LOND-D505F/2020\|EPI_ISL_444197\|202 | ···················· | ···················· | ···················· | ···················· | ···················· | ···················· |
| B | 9701 | hCoV-19/England/LOND-D508C/2020\|EPI_ISL_444198\|202 | ···················· | ···················· | ···················· | ···················· | ···················· | ···················· |
| B | 9702 | hCoV-19/England/LOND-D509B/2020\|EPI_ISL_444199\|202 | ···················· | ···················· | ···················· | ···················· | ···················· | ···················· |
| B | 9703 | hCoV-19/Wales/PHWC-28CB3/2020\|EPI_ISL_445427\|2020- | ···················· | ···················· | ···················· | ···················· | ···················· | ···················· |
| B | 9704 | hCoV-19/Wales/PHWC-28CC2/2020\|EPI_ISL_445428\|2020- | ···················· | ···················· | ···················· | ···················· | ···················· | ···················· |
| B | 9705 | hCoV-19/Wales/PHWC-28D38/2020\|EPI_ISL_445433\|2020- | ···················· | ···················· | ···················· | ···················· | ···················· | ···················· |
| B | 9706 | hCoV-19/Wales/PHWC-28D56/2020\|EPI_ISL_445434\|2020- | ···················· | ···A················ | ···················· | ···················· | ···················· | ···················· |
| B | 9707 | hCoV-19/Wales/PHWC-28D92/2020\|EPI_ISL_445438\|2020- | ···················· | ···················· | ···················· | ···················· | ···················· | ···················· |
| B | 9708 | hCoV-19/Wales/PHWC-28DA1/2020\|EPI_ISL_445439\|2020- | ···················· | ···A················ | ···················· | ···················· | ···················· | ···················· |
| B | 9709 | hCoV-19/Wales/PHWC-28E44/2020\|EPI_ISL_445448\|2020- | ···················· | ···················· | ···················· | ···················· | ···················· | ···················· |
| B | 9710 | hCoV-19/Wales/PHWC-28E80/2020\|EPI_ISL_445452\|2020- | ···················· | ···M················ | ···················· | ···················· | ···················· | ···················· |
| B | 9711 | hCoV-19/Wales/PHWC-28E9F/2020\|EPI_ISL_445453\|2020- | ···················· | ···················· | ···················· | ···················· | ···················· | ···················· |
| B | 9712 | hCoV-19/Wales/PHWC-28EAE/2020\|EPI_ISL_445454\|2020- | ···················· | ···················· | ···················· | ···················· | ···················· | ···················· |
| B | 9713 | hCoV-19/Wales/PHWC-28EDB/2020\|EPI_ISL_445457\|2020- | ···················· | ···················· | ···················· | ···················· | ···················· | ···················· |
| B | 9714 | hCoV-19/Wales/PHWC-28EEA/2020\|EPI_ISL_445458\|2020- | ···················· | ···················· | ···················· | ···················· | ···················· | ···················· |
| B | 9715 | hCoV-19/Wales/PHWC-28F23/2020\|EPI_ISL_445459\|2020- | ···················· | ···················· | ···················· | ···················· | ···················· | ···················· |
| B | 9716 | hCoV-19/Wales/PHWC-28F50/2020\|EPI_ISL_445462\|2020- | ···················· | ···················· | ···················· | ···················· | ···················· | ···················· |
| B | 9717 | hCoV-19/Wales/PHWC-291FF/2020\|EPI_ISL_445494\|2020- | ···················· | ···A················ | ···················· | ···················· | ···················· | ···················· |
| B | 9718 | hCoV-19/Wales/PHWC-2921A/2020\|EPI_ISL_445496\|2020- | ···················· | ···A················ | ···················· | ···················· | ···················· | ···················· |
| B | 9719 | hCoV-19/Wales/PHWC-33866/2020\|EPI_ISL_446758\|2020- | ···················· | ···················· | ···················· | ···················· | ···················· | ···················· |
| B | 9720 | hCoV-19/Wales/PHWC-338C0/2020\|EPI_ISL_446762\|2020- | ···················· | ···A················ | ···················· | ···················· | ···················· | ···················· |
| B | 9721 | hCoV-19/Wales/PHWC-33927/2020\|EPI_ISL_446767\|2020- | ···················· | ···················· | ···················· | ···················· | ···················· | ···················· |
| B | 9722 | hCoV-19/Wales/PHWC-33963/2020\|EPI_ISL_446771\|2020- | ···················· | ···················· | ···················· | ···················· | ···················· | ···················· |

| | | | | | | | | | |
|---|---|---|---|---|---|---|---|---|---|
| B | 9723 | hCoV-19/Wales/PHWC-339BE/2020\|EPI_ISL_446774\|2020- | ···················· | ···················· | ···················· | ···················· | ···················· | ···················· | |
| B | 9724 | hCoV-19/Wales/PHWC-33A8E/2020\|EPI_ISL_446780\|2020- | ···················· | ···················· | ···················· | ···················· | ···················· | ···················· | |
| B | 9725 | hCoV-19/Wales/PHWC-33B12/2020\|EPI_ISL_446788\|2020- | ···················· | ···················· | ···················· | ···················· | ···················· | ···················· | |
| B | 9726 | hCoV-19/Wales/PHWC-33B7C/2020\|EPI_ISL_446794\|2020- | ···················· | ···················· | ···················· | ···················· | ···················· | ···················· | |
| B | 9727 | hCoV-19/Wales/PHWC-33B8B/2020\|EPI_ISL_446795\|2020- | ···················· | ···················· | ···················· | ···················· | ···················· | ···················· | |
| B | 9728 | hCoV-19/Wales/PHWC-33B9A/2020\|EPI_ISL_446796\|2020- | ···················· | ···················· | ···················· | ···················· | ···················· | ···················· | |
| B | 9729 | hCoV-19/Wales/PHWC-2771D/2020\|EPI_ISL_432176\|2020- | ···················· | ···A················ | ···················· | ···················· | ···················· | ···················· | |
| B | 9730 | hCoV-19/Wales/PHWC-252F0/2020\|EPI_ISL_432177\|2020- | ···················· | ···················· | ···················· | ···················· | ···················· | ···················· | |
| B | 9731 | hCoV-19/Wales/PHWC-2A12B/2020\|EPI_ISL_432178\|2020- | ···················· | ···················· | ···················· | ···················· | ···················· | ···················· | |
| B | 9732 | hCoV-19/Wales/PHWC-25384/2020\|EPI_ISL_432179\|2020- | ···················· | ···················· | ···················· | ···················· | ···················· | ···················· | |
| B | 9733 | hCoV-19/Wales/PHWC-27A9C/2020\|EPI_ISL_432180\|2020- | ···················· | ·A·················· | ···················· | ···················· | ···················· | ···················· | |
| B | 9734 | hCoV-19/Wales/PHWC-29E16/2020\|EPI_ISL_432181\|2020- | ···················· | ·A·················· | ···················· | ···················· | ···················· | ···················· | |
| B | 9735 | hCoV-19/Wales/PHWC-2531B/2020\|EPI_ISL_432182\|2020- | ···················· | ···················· | ···················· | ···················· | ···················· | ···················· | |
| B | 9736 | hCoV-19/Wales/PHWC-25366/2020\|EPI_ISL_432183\|2020- | ···················· | ···················· | ···················· | ···················· | ···················· | ···················· | |
| B | 9737 | hCoV-19/Wales/PHWC-27CA5/2020\|EPI_ISL_432185\|2020- | ···················· | ···················· | ···················· | ···················· | ···················· | ···················· | |
| B | 9738 | hCoV-19/Wales/PHWC-29E61/2020\|EPI_ISL_432186\|2020- | ···················· | ···················· | ···················· | ···················· | ···················· | ···················· | |
| B | 9739 | hCoV-19/Wales/PHWC-2A06A/2020\|EPI_ISL_432187\|2020- | ···················· | ···················· | ···················· | ···················· | ···················· | ···················· | |
| B | 9740 | hCoV-19/Wales/PHWC-25393/2020\|EPI_ISL_432190\|2020- | ···················· | ···A················ | ···················· | ···················· | ···················· | ···················· | |
| B | 9741 | hCoV-19/Wales/PHWC-2B2BE/2020\|EPI_ISL_432191\|2020- | ···················· | ···················· | ···················· | ···················· | ···················· | ···················· | |
| B | 9742 | hCoV-19/Wales/PHWC-27CD2/2020\|EPI_ISL_432192\|2020- | ···················· | ···················· | ···················· | ···················· | ···················· | ···················· | |
| B | 9743 | hCoV-19/Wales/PHWC-29BC4/2020\|EPI_ISL_432193\|2020- | ···················· | ···················· | ···················· | ···················· | ···················· | ···················· | |
| B | 9744 | hCoV-19/Wales/PHWC-29E43/2020\|EPI_ISL_432194\|2020- | ···················· | ···················· | ···················· | ···················· | ···················· | ···················· | |
| B | 9745 | hCoV-19/Wales/PHWC-29C3A/2020\|EPI_ISL_432195\|2020- | ···················· | ···················· | ···················· | ···················· | ···················· | ···················· | |
| B | 9746 | hCoV-19/Wales/PHWC-29C49/2020\|EPI_ISL_432196\|2020- | ···················· | ···················· | ···················· | ···················· | ···················· | ···················· | |
| B | 9747 | hCoV-19/Wales/PHWC-29DDD/2020\|EPI_ISL_432197\|2020- | ···················· | ···················· | ···················· | ···················· | ···················· | ···················· | |
| B | 9748 | hCoV-19/Wales/PHWC-27ABA/2020\|EPI_ISL_432198\|2020- | ···················· | ···················· | ···················· | ···················· | ···················· | ···················· | |
| B | 9749 | hCoV-19/Wales/PHWC-29F13/2020\|EPI_ISL_432199\|2020- | ···················· | ···················· | ···················· | ···················· | ···················· | ···················· | |
| B | 9750 | hCoV-19/England/CAMB-7FBFC/2020\|EPI_ISL_433490\|202 | ···················· | ···················· | ···················· | ···················· | ···················· | ···················· | |
| B | 9751 | hCoV-19/England/CAMB-7FC08/2020\|EPI_ISL_433491\|202 | ···················· | ···················· | ···················· | ···················· | ···················· | ···················· | |
| B | 9752 | hCoV-19/England/CAMB-7FC17/2020\|EPI_ISL_433492\|202 | ···················· | ···················· | ···················· | ···················· | ···················· | ···················· | |
| B | 9753 | hCoV-19/England/CAMB-7FC26/2020\|EPI_ISL_433493\|202 | ···················· | ···················· | ···················· | ···················· | ···················· | ···················· | |
| B | 9754 | hCoV-19/England/CAMB-7FC35/2020\|EPI_ISL_433494\|202 | ···················· | ···················· | ···················· | ···················· | ···················· | ···················· | |
| B | 9755 | hCoV-19/England/CAMB-7FC44/2020\|EPI_ISL_433495\|202 | ···················· | ···················· | ···················· | ···················· | ···················· | ···················· | |
| B | 9756 | hCoV-19/England/CAMB-7FC53/2020\|EPI_ISL_433496\|202 | ···················· | ···················· | ···················· | ···················· | ···················· | ···················· | |
| B | 9757 | hCoV-19/England/CAMB-7FC62/2020\|EPI_ISL_433497\|202 | ···················· | ···················· | ···················· | ···················· | ···················· | ···················· | |
| B | 9758 | hCoV-19/Scotland/CVR1000/2020\|EPI_ISL_433498\|2020- | ···················· | ···················· | ···················· | ···················· | ···················· | ···················· | |
| B | 9759 | hCoV-19/Scotland/CVR1022/2020\|EPI_ISL_433499\|2020- | ···················· | ···················· | ···················· | ···················· | ···················· | ···················· | |
| B | 9760 | hCoV-19/England/LOND-D45E8/2020\|EPI_ISL_444139\|202 | ···················· | ···················· | ···················· | ···················· | ···················· | ···················· | |
| B | 9761 | hCoV-19/England/LOND-D4603/2020\|EPI_ISL_444141\|202 | ···················· | ···················· | ···················· | ···················· | ···················· | ···················· | |
| B | 9762 | hCoV-19/England/LOND-D4621/2020\|EPI_ISL_444143\|202 | ···················· | ···················· | ···················· | ···················· | ···················· | ···················· | |
| B | 9763 | hCoV-19/England/LOND-D4630/2020\|EPI_ISL_444144\|202 | ···················· | ···················· | ···················· | ···················· | ···················· | ···················· | |
| B | 9764 | hCoV-19/England/LOND-D46C7/2020\|EPI_ISL_444146\|202 | ···················· | ···················· | ···················· | ···················· | ···················· | ···················· | |
| B | 9765 | hCoV-19/England/LOND-D472E/2020\|EPI_ISL_444147\|202 | ···················· | ···················· | ···················· | ···················· | ···················· | ···················· | |
| B | 9766 | hCoV-19/England/LOND-D474C/2020\|EPI_ISL_444149\|202 | ···················· | ···················· | ···················· | ···················· | ···················· | ···················· | |
| B | 9767 | hCoV-19/England/LOND-D476A/2020\|EPI_ISL_444150\|202 | ···················· | ···················· | ···················· | ···················· | ···················· | ···················· | |
| B | 9768 | hCoV-19/England/LOND-D4779/2020\|EPI_ISL_444151\|202 | ···················· | ···················· | ···················· | ···················· | ···················· | ···················· | |
| B | 9769 | hCoV-19/England/LOND-D4788/2020\|EPI_ISL_444152\|202 | ···················· | ···················· | ···················· | ···················· | ···················· | ···················· | |
| B | 9770 | hCoV-19/England/LOND-D4797/2020\|EPI_ISL_444153\|202 | ···················· | ···················· | ···················· | ···················· | ···················· | ···················· | |
| B | 9771 | hCoV-19/England/LOND-D47D3/2020\|EPI_ISL_444155\|202 | ···················· | ···················· | ···················· | ···················· | ···················· | ···················· | |
| B | 9772 | hCoV-19/England/LOND-D4867/2020\|EPI_ISL_444156\|202 | ···················· | ···················· | ···················· | ···················· | ···················· | ···················· | |
| B | 9773 | hCoV-19/England/LOND-D4964/2020\|EPI_ISL_444159\|202 | ···················· | ···················· | ···················· | ···················· | ···················· | ···················· | |
| B | 9774 | hCoV-19/England/LOND-D4A25/2020\|EPI_ISL_444170\|202 | ···················· | ···················· | ···················· | ···················· | ···················· | ···················· | |
| B | 9775 | hCoV-19/England/LOND-D4A34/2020\|EPI_ISL_444171\|202 | ···················· | ···················· | ···················· | ···················· | ···················· | ···················· | |
| B | 9776 | hCoV-19/England/LOND-D4A52/2020\|EPI_ISL_444172\|202 | ···················· | ···················· | ···················· | ···················· | ···················· | ···················· | |
| B | 9777 | hCoV-19/England/LOND-D4A61/2020\|EPI_ISL_444173\|202 | ···················· | ···················· | ···················· | ···················· | ···················· | ···················· | |
| B | 9778 | hCoV-19/England/LOND-D4A70/2020\|EPI_ISL_444174\|202 | ···················· | ···················· | ···················· | ···················· | ···················· | ···················· | |
| B | 9779 | hCoV-19/England/LOND-D4A8F/2020\|EPI_ISL_444175\|202 | ···················· | ·A·················· | ···················· | ···················· | ···················· | ···················· | |
| B | 9780 | hCoV-19/England/LOND-D4ACB/2020\|EPI_ISL_444178\|202 | ···················· | ···················· | ···················· | ···················· | ···················· | ···················· | |
| B | 9781 | hCoV-19/England/LOND-D4AE9/2020\|EPI_ISL_444179\|202 | ···················· | ···················· | ···················· | ···················· | ···················· | ···················· | |
| B | 9782 | hCoV-19/Wales/PHWC-2902F/2020\|EPI_ISL_445470\|2020- | ···················· | ···················· | ···················· | ···················· | ···················· | ···················· | |
| B | 9783 | hCoV-19/Wales/PHWC-2907A/2020\|EPI_ISL_445475\|2020- | ···················· | ···················· | ···················· | ···················· | ···················· | ···················· | |
| B | 9784 | hCoV-19/Wales/PHWC-290B6/2020\|EPI_ISL_445478\|2020- | ···················· | ···················· | ···················· | ···················· | ···················· | ···················· | |
| B | 9785 | hCoV-19/Wales/PHWC-290F2/2020\|EPI_ISL_445481\|2020- | ···················· | ···················· | ···················· | ···················· | ···················· | ···················· | |
| B | 9786 | hCoV-19/Wales/PHWC-2913B/2020\|EPI_ISL_445485\|2020- | ···················· | ···················· | ···················· | ···················· | ···················· | ···················· | |
| B | 9787 | hCoV-19/Wales/PHWC-29177/2020\|EPI_ISL_445489\|2020- | ···················· | ···A················ | ···················· | ···················· | ···················· | ···················· | |
| C | 1 | hCoV-19/Scotland/CVR2877/2020\|EPI_ISL_448201\|2020- | ···················· | ···················· | ···················· | ···················· | ···················· | ···················· | |
| C | 2 | hCoV-19/Scotland/CVR2888/2020\|EPI_ISL_448204\|2020- | ···················· | ···················· | ···················· | ···················· | ···················· | ···················· | |
| C | 3 | hCoV-19/Scotland/CVR2893/2020\|EPI_ISL_448205\|2020- | ···················· | ·A·················· | ···················· | ···················· | ···················· | ···················· | |
| C | 4 | hCoV-19/Scotland/CVR2930/2020\|EPI_ISL_448206\|2020- | ···················· | ···················· | ···················· | ···················· | ···················· | ···················· | |
| C | 5 | hCoV-19/Scotland/CVR2932/2020\|EPI_ISL_448207\|2020- | ···················· | ···················A | ···················· | ···················· | ···················· | ···················· | 1 |
| C | 6 | hCoV-19/England/NORW-E89D6/2020\|EPI_ISL_448260\|202 | ···················· | ···················· | ···················· | ···················· | ···················· | ···················· | |
| C | 7 | hCoV-19/England/NORW-E89E5/2020\|EPI_ISL_448261\|202 | ···················· | ···················· | ···················· | ···················· | ···················· | ···················· | |
| C | 8 | hCoV-19/England/NORW-E89F4/2020\|EPI_ISL_448262\|202 | ···················· | ···················· | ···················· | ···················· | ···················· | ···················· | |
| C | 9 | hCoV-19/England/NORW-E8A00/2020\|EPI_ISL_448263\|202 | ···················· | ···················· | ···················· | ···················· | ···················· | ···················· | |
| C | 10 | hCoV-19/England/NORW-E8A1F/2020\|EPI_ISL_448264\|202 | ···················· | ···················· | ···················· | ···················· | ···················· | ···················· | |
| C | 11 | hCoV-19/England/NORW-E8A2E/2020\|EPI_ISL_448265\|202 | ···················· | ···················· | ···················· | ···················· | ···················· | ···················· | |
| C | 12 | hCoV-19/England/NORW-E8A3D/2020\|EPI_ISL_448266\|202 | ···················· | ···················· | ···················· | ···················· | ···················· | ···················· | |
| C | 13 | hCoV-19/England/NORW-E8A4C/2020\|EPI_ISL_448267\|202 | ···················· | ···················· | ···················· | ···················· | ···················· | ···················· | |

```
C 14 hCoV-19/England/NORW-E8A5B/2020|EPI_ISL_448268|202 .................... .................... .................... .................... .................... ....................
C 15 hCoV-19/England/NORW-E8A6A/2020|EPI_ISL_448269|202 .................... .................... .................... .................... .................... ....................
C 16 hCoV-19/England/NORW-E8A79/2020|EPI_ISL_448270|202 .................... .................... .................... .................... .................... ....................
C 17 hCoV-19/England/NORW-E8A88/2020|EPI_ISL_448271|202 .................... .................... .................... .................... .................... ....................
C 18 hCoV-19/England/NORW-E8A97/2020|EPI_ISL_448272|202 .................... .................... .................... .................... .................... ....................
C 19 hCoV-19/England/NORW-E8AA6/2020|EPI_ISL_448273|202 .................... .................... .................... .................... .................... ....................
C 20 hCoV-19/England/NORW-E8AB5/2020|EPI_ISL_448274|202 .................... .................... .................... .................... .................... ....................
C 21 hCoV-19/England/NORW-E8AD3/2020|EPI_ISL_448276|202 .................... .................... .................... .................... .................... ....................
C 22 hCoV-19/England/NORW-E8AE2/2020|EPI_ISL_448277|202 .................... .................... .................... .................... .................... ....................
C 23 hCoV-19/England/NORW-E8AF1/2020|EPI_ISL_448278|202 .................... .................... .................... .................... .................... ....................
C 24 hCoV-19/England/NORW-E8B0D/2020|EPI_ISL_448279|202 .................... .................... .................... .................... .................... ....................
C 25 hCoV-19/England/NORW-E8B1C/2020|EPI_ISL_448280|202 .................... .................... .................... .................... .................... ....................
C 26 hCoV-19/England/NORW-E8B3A/2020|EPI_ISL_448281|202 .................... .................... .................... .................... .................... ....................
C 27 hCoV-19/England/NORW-E8B58/2020|EPI_ISL_448282|202 .................... .................... .................... .................... .................... ....................
C 28 hCoV-19/England/NORW-E8B85/2020|EPI_ISL_448283|202 .................... .................... .................... .................... .................... ....................
C 29 hCoV-19/England/NORW-E8BA3/2020|EPI_ISL_448285|202 .................... .................... .................... .................... .................... ....................
C 30 hCoV-19/England/NORW-E8BB2/2020|EPI_ISL_448286|202 .................... .................... .................... .................... .................... ....................
C 31 hCoV-19/England/NORW-E8BC1/2020|EPI_ISL_448287|202 .................... .................... .................... .................... .................... ....................
C 32 hCoV-19/England/NORW-E8BD0/2020|EPI_ISL_448288|202 .................... .................... .................... .................... .................... ....................
C 33 hCoV-19/England/NORW-E8BEF/2020|EPI_ISL_448289|202 .................... .................... .................... .................... .................... ....................
C 34 hCoV-19/England/NORW-E8BFE/2020|EPI_ISL_448290|202 .................... .................... .................... .................... .................... ....................
C 35 hCoV-19/England/NORW-E8C0A/2020|EPI_ISL_448291|202 .................... .................... .................... .................... .................... ....................
C 36 hCoV-19/England/NORW-E8C37/2020|EPI_ISL_448292|202 .................... .................... .................... .................... .................... ....................
C 37 hCoV-19/England/NORW-E8C64/2020|EPI_ISL_448293|202 .................... .................... .................... .................... .................... ....................
C 38 hCoV-19/England/NORW-E8C73/2020|EPI_ISL_448294|202 .................... .................... .................... .................... .................... ....................
C 39 hCoV-19/England/NORW-E8C82/2020|EPI_ISL_448295|202 .................... .................... .................... .................... .................... ....................
C 40 hCoV-19/England/NORW-E8C91/2020|EPI_ISL_448296|202 .................... .................... .................... .................... .................... ....................
C 41 hCoV-19/England/NORW-E8CA0/2020|EPI_ISL_448297|202 .................... .................... .................... .................... .................... ....................
C 42 hCoV-19/England/NORW-E8CCE/2020|EPI_ISL_448299|202 .................... ...................A .................... .................... .................... .................... 1
C 43 hCoV-19/Latvia/020/2020|EPI_ISL_450520|2020-04-28 .................... .................... .................... .................... .................... ....................
C 44 hCoV-19/Latvia/021/2020|EPI_ISL_450521|2020-04-28 .................... .................... .................... .................... .................... ....................
C 45 hCoV-19/Latvia/022/2020|EPI_ISL_450522|2020-04-28 .................... .................... .................... .................... .................... ....................
C 46 hCoV-19/Latvia/023/2020|EPI_ISL_450523|2020-04-28 .................... .................... .................... .................... .................... ....................
C 47 hCoV-19/Latvia/024/2020|EPI_ISL_450524|2020-04-19 .................... .................... .................... .................... .................... ....................
C 48 hCoV-19/Poland/1109201/2020|EPI_ISL_450525|2020-03 .................... .................... .................... .................... .................... ....................
C 49 hCoV-19/USA/MI-MDHHS-SC20363/2020|EPI_ISL_450574|2 .................... .................... .................... .................... .................... ....................
C 50 hCoV-19/USA/MI-MDHHS-SC20365/2020|EPI_ISL_450576|2 .................... .................... .................... .................... .................... ....................
C 51 hCoV-19/USA/MI-MDHHS-SC20366/2020|EPI_ISL_450577|2 .................... .................... .................... .................... .................... ....................
C 52 hCoV-19/USA/MI-MDHHS-SC20369/2020|EPI_ISL_450580|2 .................... .................... .................... .................... .................... ....................
C 53 hCoV-19/USA/MI-MDHHS-SC20371/2020|EPI_ISL_450581|2 .................... .................... .................... .................... .................... ....................
C 54 hCoV-19/USA/MI-MDHHS-SC20374/2020|EPI_ISL_450585|2 .................... .................... .................... .................... .................... ....................
C 55 hCoV-19/USA/MI-MDHHS-SC20377/2020|EPI_ISL_450588|2 .................... .................... .................... .................... .................... ....................
C 56 hCoV-19/USA/MI-MDHHS-SC20381/2020|EPI_ISL_450592|2 .................... .................... .................... .................... .................... ....................
C 58 hCoV-19/Portugal/PT0384/2020|EPI_ISL_454100|2020-0 .................... .................... .................... .................... .................... ....................
C 59 hCoV-19/Portugal/PT0385/2020|EPI_ISL_454101|2020-0 .................... .................... .................... .................... .................... ....................
C 60 hCoV-19/Portugal/PT0387/2020|EPI_ISL_454103|2020-0 .................... .................... .................... .................... .................... ....................
C 61 hCoV-19/Portugal/PT0388/2020|EPI_ISL_454104|2020-0 .................... .................... .................... .................... .................... ....................
C 62 hCoV-19/Portugal/PT0389b/2020|EPI_ISL_454106|2020- .................... .................... .................... .................... .................... ....................
C 63 hCoV-19/Portugal/PT0392/2020|EPI_ISL_454111|2020-0 .................... .................... .................... .................... .................... ....................
C 64 hCoV-19/Portugal/PT0395a/2020|EPI_ISL_454114|2020- .................... .................... .................... .................... .................... ....................
C 65 hCoV-19/Portugal/PT0395b/2020|EPI_ISL_454115|2020- .................... .................... .................... .................... .................... ....................
C 66 hCoV-19/Portugal/PT0395c/2020|EPI_ISL_454116|2020- .................... .................... .................... .................... .................... ....................
C 67 hCoV-19/Portugal/PT0396b/2020|EPI_ISL_454118|2020- .................... .................... .................... .................... .................... ....................
C 68 hCoV-19/Portugal/PT0399a/2020|EPI_ISL_454122|2020- .................... .................... .................... .................... .................... ....................
C 69 hCoV-19/Portugal/PT0399b/2020|EPI_ISL_454123|2020- .................... .................... .................... .................... .................... ....................
C 70 hCoV-19/Portugal/PT0403/2020|EPI_ISL_454127|2020-0 .................... .................... .................... .................... .................... ....................
C 71 hCoV-19/Portugal/PT0404/2020|EPI_ISL_454128|2020-0 .................... .................... .................... .................... .................... ....................
C 72 hCoV-19/Portugal/PT0405/2020|EPI_ISL_454129|2020-0 .................... .................... .................... .................... .................... ....................
C 73 hCoV-19/Portugal/PT0406/2020|EPI_ISL_454130|2020-0 .................... .................... .................... .................... .................... ....................
C 74 hCoV-19/Portugal/PT0407/2020|EPI_ISL_454131|2020-0 .................... .................... .................... .................... .................... ....................
C 75 hCoV-19/Portugal/PT0408/2020|EPI_ISL_454132|2020-0 .................... .................... .................... .................... .................... ....................
C 76 hCoV-19/Portugal/PT0409/2020|EPI_ISL_454133|2020-0 .................... .................... .................... .................... .................... ....................
C 77 hCoV-19/Portugal/PT0410/2020|EPI_ISL_454134|2020-0 .................... .................... .................... .................... .................... ....................
C 78 hCoV-19/Portugal/PT0411/2020|EPI_ISL_454135|2020-0 .................... .................... .................... .................... .................... ....................
C 79 hCoV-19/Wuhan/HB-WHCM-118/2020|EPI_ISL_455406|2020 .................... .................... .................... .................... .................... ....................
C 80 hCoV-19/Nigeria/OG007-CV22/2020|EPI_ISL_455412|202 .................... .................... .................... .................... .................... ....................
C 82 hCoV-19/Nigeria/OS060-CV9/2020|EPI_ISL_455419|2020 .................... .................... .................... .................... .................... ....................
C 83 hCoV-19/Bangladesh/BCSIR-NILMRC-007/2020|EPI_ISL_4 .................... .................... .................... .................... .................... ....................
C 84 hCoV-19/Nigeria/OS070-CV11/2020|EPI_ISL_455422|202 .................... .................... .................... .................... .................... ....................
C 85 hCoV-19/Nigeria/OS075-CV12/2020|EPI_ISL_455423|202 .................... .................... .................... .................... .................... ....................
C 86 hCoV-19/Nigeria/OS085-CV14/2020|EPI_ISL_455424|202 .................... .................... .................... .................... .................... ....................
C 87 hCoV-19/Nigeria/OS122-CV18/2020|EPI_ISL_455426|202 .................... .................... .................... .................... .................... ....................
C 88 hCoV-19/Nigeria/OY008-CV29/2020|EPI_ISL_455429|202 .................... .................... .................... .................... .................... ....................
C 89 hCoV-19/Nigeria/OY045A-CV35/2020|EPI_ISL_455431|20 .................... .................... .................... .................... .................... ....................
C 90 hCoV-19/Mexico/GRO-InDRE-10/2020|EPI_ISL_455434|20 .................... .................... .................... .................... .................... ....................
C 91 hCoV-19/Mexico/JAL-InDRE-11/2020|EPI_ISL_455435|20 .................... .................... .................... .................... .................... ....................
C 93 hCoV-19/Mexico/CMX-InDRE-13/2020|EPI_ISL_455437|20 .................... .................... .................... .................... .................... ....................
C 94 hCoV-19/Mexico/ROO-InDRE-14/2020|EPI_ISL_455438|20 .................... .................... .................... .................... .................... ....................
```

| | | | | | | | | | |
|---|---|---|---|---|---|---|---|---|---|
| C | 95 | hCoV-19/Mexico/CMX-InDRE_15/2020\|EPI_ISL_455439\|20 | .................... | .................... | .................... | .................... | .................... | .................... | |
| C | 96 | hCoV-19/Poland/PL_P27/2020\|EPI_ISL_455440\|2020-03- | .................... | .................... | .................... | .................... | .................... | .................... | |
| C | 97 | hCoV-19/Poland/PL_P28/2020\|EPI_ISL_455441\|2020-04- | .................... | .................... | .................... | .................... | .................... | .................... | |
| C | 98 | hCoV-19/Poland/PL_P29/2020\|EPI_ISL_455442\|2020-03- | .................... | .................... | .................... | .................... | .................... | .................... | |
| C | 99 | hCoV-19/Poland/PL_P30/2020\|EPI_ISL_455443\|2020-04- | .................... | .................... | .................... | .................... | .................... | .................... | |
| C | 100 | hCoV-19/Poland/PL_P31/2020\|EPI_ISL_455444\|2020-03- | .................... | .................... | .................... | .................... | .................... | .................... | |
| C | 101 | hCoV-19/Poland/PL_P32/2020\|EPI_ISL_455445\|2020-03- | .................... | .................... | .................... | .................... | .................... | .................... | |
| C | 102 | hCoV-19/Poland/PL_P33/2020\|EPI_ISL_455446\|2020-03- | .................... | .................... | .................... | .................... | .................... | .................... | |
| C | 103 | hCoV-19/Poland/PL_P34/2020\|EPI_ISL_455447\|2020-03- | .................... | .................... | .................... | .................... | .................... | .................... | |
| C | 104 | hCoV-19/Poland/PL_P35/2020\|EPI_ISL_455448\|2020-04- | .................... | .................... | .................... | .................... | .................... | .................... | |
| C | 105 | hCoV-19/Poland/PL_P36/2020\|EPI_ISL_455449\|2020-03- | .................... | .................... | .................... | .................... | .................... | .................... | |
| C | 106 | hCoV-19/Poland/PL_P37/2020\|EPI_ISL_455450\|2020-03- | .................... | .................... | .................... | .................... | .................... | .................... | |
| C | 107 | hCoV-19/Poland/PL_P38/2020\|EPI_ISL_455451\|2020-03- | .................... | .................... | .................... | .................... | .................... | .................... | |
| C | 108 | hCoV-19/Poland/PL_P39/2020\|EPI_ISL_455452\|2020-03- | .................... | .................... | .................... | .................... | .................... | .................... | |
| C | 109 | hCoV-19/Poland/PL_P40/2020\|EPI_ISL_455453\|2020-03- | .................... | .................... | .................... | .................... | .................... | .................... | |
| C | 110 | hCoV-19/Mexico/PUE-InDRE-17/2020\|EPI_ISL_455455\|20 | .................... | .................... | .................... | .................... | .................... | .................... | |
| C | 111 | hCoV-19/Mexico/QUE-InDRE-18/2020\|EPI_ISL_455456\|20 | .................... | .................... | .................... | .................... | .................... | .................... | |
| C | 112 | hCoV-19/Bangladesh/BCSIR-NILMRC-006/2020\|EPI_ISL_4 | .................... | .................... | .................... | .................... | .................... | .................... | |
| C | 113 | hCoV-19/Bangladesh/BCSIR-NILMRC-008/2020\|EPI_ISL_4 | .................... | .................... | .................... | .................... | .................... | .................... | |
| C | 114 | hCoV-19/Yichun/JX2325/2020\|EPI_ISL_455460\|2020-02- | .................... | .................... | .................... | .................... | .................... | .................... | |
| C | 115 | hCoV-19/Yichun/JX2391/2020\|EPI_ISL_455461\|2020-02- | .................... | .................... | .................... | .................... | .................... | .................... | |
| C | 116 | hCoV-19/Fuzhou/JX2012/2020\|EPI_ISL_455462\|2020-02- | .................... | .................... | .................... | .................... | .................... | .................... | |
| C | 117 | hCoV-19/Jiujiang/JX490/2020\|EPI_ISL_455463\|2020-02 | .................... | .................... | .................... | .................... | .................... | .................... | |
| C | 118 | hCoV-19/Fuzhou/JX2009/2020\|EPI_ISL_455464\|2020-02- | .................... | .................... | .................... | .................... | .................... | .................... | |
| C | 119 | hCoV-19/Fuzhou/JX2019/2020\|EPI_ISL_455465\|2020-02- | .................... | .................... | .................... | .................... | .................... | .................... | |
| C | 120 | hCoV-19/Yingtan/JX2480/2020\|EPI_ISL_455466\|2020-02 | .................... | .................... | .................... | .................... | .................... | .................... | |
| C | 121 | hCoV-19/Yichun/JX2424/2020\|EPI_ISL_455467\|2020-02- | .................... | .................... | .................... | .................... | .................... | .................... | |
| C | 122 | hCoV-19/England/CAMB-1B2673/2020\|EPI_ISL_456702\|20 | .................... | .................... | .................... | .................... | .................... | .................... | |
| C | 123 | hCoV-19/England/CAMB-1B2716/2020\|EPI_ISL_456704\|20 | .................... | .................... | .................... | .................... | .................... | .................... | |
| C | 124 | hCoV-19/England/CAMB-1B2901/2020\|EPI_ISL_456705\|20 | .................... | .................... | .................... | .................... | .................... | .................... | |
| C | 125 | hCoV-19/England/CAMB-1B2B92/2020\|EPI_ISL_456707\|20 | .................... | .................... | .................... | .................... | .................... | .................... | |
| C | 126 | hCoV-19/England/CAMB-1B2BA1/2020\|EPI_ISL_456708\|20 | .................... | .................... | .................... | .................... | .................... | .................... | |
| C | 127 | hCoV-19/England/CAMB-1B2BCF/2020\|EPI_ISL_456709\|20 | .................... | .................... | .................... | .................... | .................... | .................... | |
| C | 128 | hCoV-19/England/CAMB-1B2BED/2020\|EPI_ISL_456711\|20 | .................... | .................... | .................... | .................... | .................... | .................... | |
| C | 129 | hCoV-19/England/CAMB-1B2C08/2020\|EPI_ISL_456713\|20 | .................... | .................... | .................... | .................... | .................... | .................... | |
| C | 130 | hCoV-19/England/CAMB-1B2C71/2020\|EPI_ISL_456716\|20 | .................... | .................... | .................... | .................... | .................... | .................... | |
| C | 131 | hCoV-19/England/CAMB-1B2CBD/2020\|EPI_ISL_456717\|20 | .................... | .................... | .................... | .................... | .................... | .................... | |
| C | 132 | hCoV-19/England/CAMB-1B2CCC/2020\|EPI_ISL_456718\|20 | .................... | .................... | .................... | .................... | .................... | .................... | |
| C | 133 | hCoV-19/England/CAMB-1B2CDB/2020\|EPI_ISL_456719\|20 | .................... | .................... | .................... | .................... | .................... | .................... | |
| C | 134 | hCoV-19/England/CAMB-1B2CF9/2020\|EPI_ISL_456720\|20 | .................... | .................... | .................... | .................... | .................... | .................... | |
| C | 135 | hCoV-19/England/CAMB-1B2D32/2020\|EPI_ISL_456721\|20 | .................... | .................... | .................... | .................... | .................... | .................... | |
| C | 136 | hCoV-19/England/CAMB-1B2D50/2020\|EPI_ISL_456722\|20 | .................... | .................... | .................... | .................... | .................... | .................... | |
| C | 137 | hCoV-19/England/CAMB-1B2D7E/2020\|EPI_ISL_456723\|20 | .................... | .................... | .................... | .................... | .................... | .................... | |
| C | 138 | hCoV-19/England/CAMB-1B2D8D/2020\|EPI_ISL_456724\|20 | .................... | .................... | .................... | .................... | .................... | .................... | |
| C | 139 | hCoV-19/England/CAMB-1B2E4E/2020\|EPI_ISL_456727\|20 | .................... | .................... | .................... | .................... | .................... | .................... | |
| C | 140 | hCoV-19/England/CAMB-1B2E7B/2020\|EPI_ISL_456728\|20 | .................... | .................... | .................... | .................... | .................... | .................... | |
| C | 141 | hCoV-19/England/CAMB-1B2E8A/2020\|EPI_ISL_456729\|20 | .................... | .................... | .................... | .................... | .................... | .................... | |
| C | 142 | hCoV-19/England/CAMB-1B2E99/2020\|EPI_ISL_456730\|20 | .................... | .................... | .................... | .................... | .................... | .................... | |
| C | 143 | hCoV-19/England/CAMB-1B2EB7/2020\|EPI_ISL_456731\|20 | .................... | .................... | .................... | .................... | .................... | .................... | |
| C | 144 | hCoV-19/England/CAMB-1B2EC6/2020\|EPI_ISL_456732\|20 | .................... | .................... | .................... | .................... | .................... | ...........A........ | |
| C | 145 | hCoV-19/England/CAMB-1B2ED5/2020\|EPI_ISL_456733\|20 | .................... | .................... | .................... | .................... | .................... | .................... | |
| C | 146 | hCoV-19/England/CAMB-1B2EE4/2020\|EPI_ISL_456734\|20 | .................... | .................... | .................... | .................... | .................... | .................... | |
| C | 147 | hCoV-19/England/CAMB-1B2F1E/2020\|EPI_ISL_456736\|20 | .................... | .................... | .................... | .................... | .................... | .................... | |
| C | 148 | hCoV-19/England/CAMB-1B2F78/2020\|EPI_ISL_456739\|20 | .................... | .................... | .................... | .................... | .................... | .................... | |
| C | 149 | hCoV-19/England/CAMB-1B2F96/2020\|EPI_ISL_456740\|20 | .................... | .................... | .................... | .................... | .................... | .................... | |
| C | 150 | hCoV-19/England/CAMB-1B2FE1/2020\|EPI_ISL_456743\|20 | .................... | .................... | .................... | .................... | .................... | .................... | |
| C | 151 | hCoV-19/England/CAMB-1B300B/2020\|EPI_ISL_456745\|20 | .................... | .................... | .................... | .................... | .................... | .................... | |
| C | 152 | hCoV-19/England/CAMB-1B3029/2020\|EPI_ISL_456747\|20 | .................... | .................... | .................... | .................... | .................... | .................... | |
| C | 153 | hCoV-19/England/CAMB-1B3047/2020\|EPI_ISL_456749\|20 | .................... | .................... | .................... | .................... | .................... | .................... | |
| C | 154 | hCoV-19/England/CAMB-1B3065/2020\|EPI_ISL_456750\|20 | .................... | .................... | .................... | .................... | .................... | .................... | |
| C | 156 | hCoV-19/England/CAMB-1B3232/2020\|EPI_ISL_456754\|20 | .................... | .................... | .................... | .................... | .................... | .................... | |
| C | 157 | hCoV-19/England/CAMB-1B3250/2020\|EPI_ISL_456755\|20 | .................... | .................... | .................... | .................... | .................... | .................... | |
| C | 160 | hCoV-19/Scotland/CVR1893/2020\|EPI_ISL_456758\|2020- | .................... | .................... | .................... | .................... | .................... | .................... | |
| C | 161 | hCoV-19/Scotland/CVR2118/2020\|EPI_ISL_456759\|2020- | .................... | .................... | .................... | .................... | .................... | .................... | |
| C | 162 | hCoV-19/Scotland/CVR3149/2020\|EPI_ISL_456766\|2020- | .................... | .................... | .................... | .................... | .................... | .................... | |
| C | 163 | hCoV-19/Scotland/CVR3167/2020\|EPI_ISL_456767\|2020- | .................... | .................... | .................... | .................... | .................... | .................... | |
| C | 164 | hCoV-19/Scotland/CVR3266/2020\|EPI_ISL_456771\|2020- | .................... | .................... | .................... | .................... | .................... | .................... | |
| C | 165 | hCoV-19/Scotland/CVR3273/2020\|EPI_ISL_456773\|2020- | .................... | .................... | .................... | .................... | .................... | .................... | |
| C | 167 | hCoV-19/Scotland/CVR3282/2020\|EPI_ISL_456776\|2020- | .................... | .................... | .................... | .................... | .................... | .................... | |
| C | 168 | hCoV-19/Scotland/CVR3284/2020\|EPI_ISL_456777\|2020- | .................... | .................... | .................... | .................... | .................... | .................... | |
| C | 169 | hCoV-19/Scotland/CVR3294/2020\|EPI_ISL_456778\|2020- | .................... | .................... | .................... | .................... | .................... | .................... | |
| C | 171 | hCoV-19/Scotland/CVR3314/2020\|EPI_ISL_456780\|2020- | .................... | .................... | .................... | .................... | .................... | .................... | |
| C | 172 | hCoV-19/Scotland/CVR3335/2020\|EPI_ISL_456781\|2020- | .................... | .A.................. | .................... | .................... | .................... | .................... | |
| C | 173 | hCoV-19/Scotland/CVR3346/2020\|EPI_ISL_456782\|2020- | .................... | .................... | .................... | .................... | .................... | .................... | |
| C | 175 | hCoV-19/Scotland/CVR3350/2020\|EPI_ISL_456784\|2020- | .................... | .................... | .................... | .................... | .................... | .................... | |
| C | 176 | hCoV-19/Scotland/CVR3353/2020\|EPI_ISL_456785\|2020- | .................... | .................... | .................... | .................... | .................... | .................... | |
| C | 179 | hCoV-19/Scotland/CVR3358/2020\|EPI_ISL_456788\|2020- | .................... | ...................A | .................... | .................... | .................... | .................... | 1 |
| C | 180 | hCoV-19/Scotland/CVR3359/2020\|EPI_ISL_456789\|2020- | .................... | ...................A | .................... | .................... | .................... | .................... | 1 |

| | | | | | | | | |
|---|---|---|---|---|---|---|---|---|
| C | 181 hCoV-19/Scotland/CVR3360/2020\|EPI_ISL_456790\|2020- | ···················· | ···················A | ···················· | ···················· | ···················· | ···················· | 1 |
| C | 182 hCoV-19/Scotland/CVR3361/2020\|EPI_ISL_456791\|2020- | ···················· | ···················· | ···················· | ···················· | ···················· | ···················· | |
| C | 183 hCoV-19/Scotland/CVR3362/2020\|EPI_ISL_456792\|2020- | ···················· | ···················· | ···················· | ···················· | ···················· | ···················· | |
| C | 184 hCoV-19/Scotland/CVR3363/2020\|EPI_ISL_456793\|2020- | ···················· | ···················A | ···················· | ···················· | ···················· | ···················· | 1 |
| C | 186 hCoV-19/Scotland/CVR3365/2020\|EPI_ISL_456795\|2020- | ···················· | ···················A | ···················· | ···················· | ···················· | ···················· | 1 |
| C | 189 hCoV-19/Scotland/CVR3368/2020\|EPI_ISL_456798\|2020- | ···················· | ···················· | ···················· | ···················· | ···················· | ···················· | |
| C | 190 hCoV-19/Netherlands/NoordHolland_32/2020\|EPI_ISL_4 | ···················· | ···················· | ···················· | ···················· | ···················· | ···················· | |
| C | 191 hCoV-19/Netherlands/NoordHolland_33/2020\|EPI_ISL_4 | ···················· | ···················· | ···················· | ···················· | ···················· | ···················· | |
| C | 192 hCoV-19/Netherlands/NoordHolland_34/2020\|EPI_ISL_4 | ···················· | ···················· | ···················· | ···················· | ···················· | ···················· | |
| C | 193 hCoV-19/Netherlands/NoordHolland_35/2020\|EPI_ISL_4 | ···················· | ···················· | ···················· | ···················· | ···················· | ···················· | |
| C | 194 hCoV-19/Netherlands/Overijssel_34/2020\|EPI_ISL_461 | ···················· | ···················· | ···················· | ···················· | ···················· | ···················· | |
| C | 195 hCoV-19/Netherlands/Overijssel_35/2020\|EPI_ISL_461 | ···················· | ···················· | ···················· | ···················· | ···················· | ···················· | |
| C | 196 hCoV-19/Netherlands/Utrecht_30/2020\|EPI_ISL_461228 | ···················· | ···················· | ···················· | ···················· | ···················· | ···················· | |
| C | 197 hCoV-19/Netherlands/Utrecht_31/2020\|EPI_ISL_461229 | ···················· | ···················· | ···················· | ···················· | ···················· | ···················· | |
| C | 198 hCoV-19/Netherlands/Utrecht_32/2020\|EPI_ISL_461230 | ···················· | ···················· | ···················· | ···················· | ···················· | ···················· | |
| C | 199 hCoV-19/Netherlands/Utrecht_34/2020\|EPI_ISL_461232 | ···················· | ···················· | ···················· | ···················· | ···················· | ···················· | |
| C | 200 hCoV-19/Netherlands/Utrecht_35/2020\|EPI_ISL_461233 | ···················· | ···················· | ···················· | ···················· | ···················· | ···················· | |
| C | 201 hCoV-19/Netherlands/Utrecht_36/2020\|EPI_ISL_461234 | ···················· | ···················· | ···················· | ···················· | ···················· | ···················· | |
| C | 202 hCoV-19/Netherlands/Utrecht_37/2020\|EPI_ISL_461235 | ···················· | ···················· | ···················· | ···················· | ···················· | ···················· | |
| C | 203 hCoV-19/Netherlands/Utrecht_38/2020\|EPI_ISL_461236 | ···················· | ···················· | ···················· | ···················· | ···················· | ···················· | |
| C | 204 hCoV-19/Netherlands/Utrecht_39/2020\|EPI_ISL_461237 | ···················· | ···················· | ···················· | ···················· | ···················· | ···················· | |
| C | 205 hCoV-19/Netherlands/Utrecht_40/2020\|EPI_ISL_461238 | ···················· | ···················· | ···················· | ···················· | ···················· | ···················· | |
| C | 206 hCoV-19/Netherlands/Utrecht_41/2020\|EPI_ISL_461239 | ···················· | ···················· | ···················· | ···················· | ···················· | ···················· | |
| C | 207 hCoV-19/Netherlands/Utrecht_42/2020\|EPI_ISL_461240 | ···················· | ···················· | ···················· | ···················· | ···················· | ···················· | |
| C | 208 hCoV-19/Netherlands/Utrecht_43/2020\|EPI_ISL_461241 | ···················· | ···················· | ···················· | ···················· | ···················· | ···················· | |
| C | 209 hCoV-19/Netherlands/Utrecht_44/2020\|EPI_ISL_461242 | ···················· | ···················· | ···················· | ···················· | ···················· | ···················· | |
| C | 210 hCoV-19/Netherlands/Utrecht_45/2020\|EPI_ISL_461243 | ···················· | ···················· | ···················· | ···················· | ···················· | ···················· | |
| C | 211 hCoV-19/Netherlands/Utrecht_48/2020\|EPI_ISL_461244 | ···················· | ···················· | ···················· | ···················· | ···················· | ···················· | |
| C | 212 hCoV-19/Netherlands/Utrecht_49/2020\|EPI_ISL_461245 | ···················· | ···················· | ···················· | ···················· | ···················· | ···················· | |
| C | 213 hCoV-19/Netherlands/Utrecht_50/2020\|EPI_ISL_461246 | ···················· | ···················· | ···················· | ···················· | ···················· | ···················· | |
| C | 214 hCoV-19/Netherlands/Zeeland_1/2020\|EPI_ISL_461247\| | ···················· | ···················· | ···················· | ···················· | ···················· | ···················· | |
| C | 215 hCoV-19/Netherlands/Zeeland_12/2020\|EPI_ISL_461250 | ···················· | ···················· | ···················· | ···················· | ···················· | ···················· | |
| C | 216 hCoV-19/Netherlands/Zeeland_13/2020\|EPI_ISL_461251 | ···················· | ···················· | ···················· | ···················· | ···················· | ···················· | |
| C | 217 hCoV-19/Netherlands/Zeeland_16/2020\|EPI_ISL_461254 | ···················· | ···················· | ···················· | ···················· | ···················· | ···················· | |
| C | 218 hCoV-19/Netherlands/Zeeland_17/2020\|EPI_ISL_461255 | ···················· | ···················· | ···················· | ···················· | ···················· | ···················· | |
| C | 219 hCoV-19/Netherlands/Zeeland_19/2020\|EPI_ISL_461257 | ···················· | ···················· | ···················· | ···················· | ···················· | ···················· | |
| C | 220 hCoV-19/Netherlands/Zeeland_2/2020\|EPI_ISL_461258\| | ···················· | ···················· | ···················· | ···················· | ···················· | ···················· | |
| C | 221 hCoV-19/Netherlands/Zeeland_20/2020\|EPI_ISL_461259 | ···················· | ···················· | ···················· | ···················· | ···················· | ···················· | |
| C | 222 hCoV-19/Netherlands/Zeeland_21/2020\|EPI_ISL_461260 | ···················· | ···················· | ···················· | ···················· | ···················· | ···················· | |
| C | 223 hCoV-19/Netherlands/Zeeland_22/2020\|EPI_ISL_461261 | ···················· | ···················· | ···················· | ···················· | ···················· | ···················· | |
| C | 224 hCoV-19/Netherlands/Zeeland_26/2020\|EPI_ISL_461264 | ···················· | ···················· | ···················· | ···················· | ···················· | ···················· | |
| C | 225 hCoV-19/Netherlands/Zeeland_27/2020\|EPI_ISL_461265 | ···················· | ···················· | ···················· | ···················· | ···················· | ···················· | |
| C | 226 hCoV-19/Netherlands/Zeeland_28/2020\|EPI_ISL_461266 | ···················· | ···················· | ···················· | ···················· | ···················· | ···················· | |
| C | 227 hCoV-19/Netherlands/Zeeland_3/2020\|EPI_ISL_461268\| | ···················· | ···················· | ···················· | ···················· | ···················· | ···················· | |
| C | 228 hCoV-19/Netherlands/Zeeland_31/2020\|EPI_ISL_461270 | ···················· | ···················· | ···················· | ···················· | ···················· | ···················· | |
| C | 229 hCoV-19/Netherlands/Zeeland_32/2020\|EPI_ISL_461271 | ···················· | ···················· | ···················· | ···················· | ···················· | ···················· | |
| C | 230 hCoV-19/Netherlands/Zeeland_33/2020\|EPI_ISL_461272 | ···················· | ···················· | ···················· | ···················· | ···················· | ···················· | |
| C | 231 hCoV-19/Netherlands/Zeeland_34/2020\|EPI_ISL_461273 | ···················· | ···················· | ···················· | ···················· | ···················· | ···················· | |
| C | 232 hCoV-19/Netherlands/Zeeland_35/2020\|EPI_ISL_461274 | ···················· | ···················· | ···················· | ···················· | ···················· | ···················· | |
| C | 233 hCoV-19/Netherlands/Zeeland_36/2020\|EPI_ISL_461275 | ···················· | ···················· | ···················· | ···················· | ···················· | ···················· | |
| C | 234 hCoV-19/Netherlands/Zeeland_37/2020\|EPI_ISL_461276 | ···················· | ···················· | ···················· | ···················· | ···················· | ···················· | |
| C | 235 hCoV-19/Netherlands/Zeeland_38/2020\|EPI_ISL_461277 | ···················· | ···················· | ···················· | ···················· | ···················· | ···················· | |
| C | 236 hCoV-19/Netherlands/Zeeland_41/2020\|EPI_ISL_461280 | ···················· | ···················· | ···················· | ···················· | ···················· | ···················· | |
| C | 237 hCoV-19/Netherlands/Zeeland_42/2020\|EPI_ISL_461281 | ···················· | ···················· | ···················· | ···················· | ···················· | ···················· | |
| C | 238 hCoV-19/Netherlands/Zeeland_43/2020\|EPI_ISL_461282 | ···················· | ···················· | ···················· | ···················· | ···················· | ···················· | |
| C | 239 hCoV-19/Netherlands/Zeeland_44/2020\|EPI_ISL_461283 | ···················· | ···················· | ···················· | ···················· | ···················· | ···················· | |
| C | 240 hCoV-19/Netherlands/Zeeland_45/2020\|EPI_ISL_461284 | ···················· | ···················· | ···················· | ···················· | ···················· | ···················· | |
| C | 241 hCoV-19/Netherlands/Zeeland_46/2020\|EPI_ISL_461285 | ···················· | ···················· | ···················· | ···················· | ···················· | ···················· | |
| C | 242 hCoV-19/Netherlands/Zeeland_47/2020\|EPI_ISL_461286 | ···················· | ···················· | ···················· | ···················· | ···················· | ···················· | |
| C | 243 hCoV-19/Netherlands/Zeeland_5/2020\|EPI_ISL_461287\| | ···················· | ···················· | ···················· | ···················· | ···················· | ···················· | |
| C | 244 hCoV-19/Netherlands/Zeeland_6/2020\|EPI_ISL_461288\| | ···················· | ···················· | ···················· | ···················· | ···················· | ···················· | |
| C | 245 hCoV-19/Netherlands/Zeeland_7/2020\|EPI_ISL_461289\| | ···················· | ···················· | ···················· | ···················· | ···················· | ···················· | |
| C | 246 hCoV-19/Netherlands/Zeeland_9/2020\|EPI_ISL_461291\| | ···················· | ···················· | ···················· | ···················· | ···················· | ···················· | |
| C | 247 hCoV-19/Netherlands/ZuidHolland_119/2020\|EPI_ISL_4 | ···················· | ···················· | ···················· | ···················· | ···················· | ···················· | |
| C | 248 hCoV-19/Netherlands/ZuidHolland_120/2020\|EPI_ISL_4 | ···················· | ···················· | ···················· | ···················· | ···················· | ···················· | |
| C | 249 hCoV-19/Netherlands/ZuidHolland_121/2020\|EPI_ISL_4 | ···················· | ···················· | ···················· | ···················· | ···················· | ···················· | |
| C | 250 hCoV-19/Netherlands/ZuidHolland_122/2020\|EPI_ISL_4 | ···················· | ···················· | ···················· | ···················· | ···················· | ···················· | |
| C | 251 hCoV-19/Netherlands/ZuidHolland_123/2020\|EPI_ISL_4 | ···················· | ···················· | ···················· | ···················· | ···················· | ···················· | |
| C | 252 hCoV-19/Netherlands/ZuidHolland_124/2020\|EPI_ISL_4 | ···················· | ···················· | ···················· | ···················· | ···················· | ···················· | |
| C | 253 hCoV-19/Netherlands/ZuidHolland_125/2020\|EPI_ISL_4 | ···················· | ···········A········ | ···················· | ···················· | ···················· | ···················· | |
| C | 254 hCoV-19/Netherlands/ZuidHolland_126/2020\|EPI_ISL_4 | ···················· | ···················· | ···················· | ···················· | ···················· | ···················· | |
| C | 255 hCoV-19/Japan/TKYE6938/2020\|EPI_ISL_468724\|2020-02 | ···················· | ···················· | ···················· | ···················· | ···················· | ···················· | |
| C | 256 hCoV-19/Japan/TKYE63442/2020\|EPI_ISL_468725\|2020-0 | ···················· | ···················· | ···················· | ···················· | ···················· | ···················· | |
| C | 257 hCoV-19/Scotland/CVR2860/2020\|EPI_ISL_448200\|2020- | ···················· | ·A·················· | ···················· | ···················· | ···················· | ···················· | |
| C | 258 hCoV-19/Scotland/CVR2944/2020\|EPI_ISL_448209\|2020- | ···················· | ···················· | ···················· | ···················· | ···················· | ···················· | |
| C | 259 hCoV-19/Scotland/CVR2970/2020\|EPI_ISL_448210\|2020- | ···················· | ···················· | ···················· | ···················· | ···················· | ···················· | |
| C | 260 hCoV-19/Scotland/CVR2971/2020\|EPI_ISL_448211\|2020- | ···················· | ···················· | ···················· | ···················· | ···················· | ···················· | |
| C | 261 hCoV-19/Scotland/CVR2972/2020\|EPI_ISL_448212\|2020- | ···················· | ···················· | ···················· | ···················· | ···················· | ···················· | |

| | | | | | | | | |
|---|---|---|---|---|---|---|---|---|
| C | 262 hCoV-19/Scotland/CVR2973/2020\|EPI_ISL_448213\|2020- | ···················· | ···················· | ···················· | ···················· | ···················· | ···················· | |
| C | 263 hCoV-19/Scotland/CVR2974/2020\|EPI_ISL_448214\|2020- | ···················· | ···················· | ···················· | ···················· | ···················· | ···················· | |
| C | 264 hCoV-19/Scotland/CVR2975/2020\|EPI_ISL_448215\|2020- | ···················· | ···················· | ···················· | ···················· | ···················· | ···················· | |
| C | 265 hCoV-19/Scotland/CVR2976/2020\|EPI_ISL_448216\|2020- | ···················· | ···················· | ···················· | ···················· | ···················· | ···················· | |
| C | 266 hCoV-19/Scotland/CVR2977/2020\|EPI_ISL_448217\|2020- | ···················· | ···················· | ···················· | ···················· | ···················· | ···················· | |
| C | 267 hCoV-19/Scotland/CVR2978/2020\|EPI_ISL_448218\|2020- | ···················· | ·A·················· | ···················· | ···················· | ···················· | ···················· | |
| C | 268 hCoV-19/Scotland/CVR2979/2020\|EPI_ISL_448219\|2020- | ···················· | ···················A | ···················· | ···················· | ···················· | ···················· | 1 |
| C | 269 hCoV-19/Scotland/CVR2980/2020\|EPI_ISL_448220\|2020- | ···················· | ···················· | ···················· | ···················· | ···················· | ···················· | |
| C | 270 hCoV-19/Scotland/CVR2981/2020\|EPI_ISL_448221\|2020- | ···················· | ···················· | ···················· | ···················· | ···················· | ···················· | |
| C | 271 hCoV-19/Vietnam/IBT-14Mar/2020\|EPI_ISL_448222\|2020 | ···················· | ···················· | ···················· | ···················· | ···················· | ···················· | |
| C | 272 hCoV-19/England/NORW-E86C0/2020\|EPI_ISL_448223\|202 | ···················· | ···················· | ···················· | ···················· | ···················· | ···················· | |
| C | 273 hCoV-19/England/NORW-E86DF/2020\|EPI_ISL_448224\|202 | ···················· | ···················· | ···················· | ···················· | ···················· | ···················· | |
| C | 274 hCoV-19/England/NORW-E86FD/2020\|EPI_ISL_448225\|202 | ···················· | ···················· | ···················· | ···················· | ···················· | ···················· | |
| C | 276 hCoV-19/England/NORW-E8718/2020\|EPI_ISL_448227\|202 | ···················· | ···················· | ···················· | ···················· | ···················· | ···················· | |
| C | 277 hCoV-19/England/NORW-E8727/2020\|EPI_ISL_448228\|202 | ···················· | ···················· | ···················· | ···················· | ···················· | ···················· | |
| C | 278 hCoV-19/England/NORW-E8745/2020\|EPI_ISL_448229\|202 | ···················· | ···················· | ···················· | ···················· | ···················· | ···················· | |
| C | 279 hCoV-19/England/NORW-E8772/2020\|EPI_ISL_448230\|202 | ···················· | ···················· | ···················· | ···················· | ···················· | ···················· | |
| C | 280 hCoV-19/England/NORW-E8790/2020\|EPI_ISL_448232\|202 | ···················· | ···················· | ···················· | ···················· | ···················· | ···················· | |
| C | 281 hCoV-19/England/NORW-E87BE/2020\|EPI_ISL_448233\|202 | ···················· | ···················· | ···················· | ···················· | ···················· | ···················· | |
| C | 282 hCoV-19/England/NORW-E87CD/2020\|EPI_ISL_448234\|202 | ···················· | ···················· | ···················· | ···················· | ···················· | ···················· | |
| C | 283 hCoV-19/England/NORW-E87DC/2020\|EPI_ISL_448235\|202 | ···················· | ···················· | ···················· | ···················· | ···················· | ···················· | |
| C | 284 hCoV-19/England/NORW-E87EB/2020\|EPI_ISL_448236\|202 | ···················· | ···················· | ···················· | ···················· | ···················· | ···················· | |
| C | 285 hCoV-19/England/NORW-E87FA/2020\|EPI_ISL_448237\|202 | ···················· | ···················· | ···················· | ···················· | ···················· | ···················· | |
| C | 286 hCoV-19/England/NORW-E8824/2020\|EPI_ISL_448238\|202 | ···················· | ···················· | ···················· | ···················· | ···················· | ···················· | |
| C | 287 hCoV-19/England/NORW-E8833/2020\|EPI_ISL_448239\|202 | ···················· | ···················· | ···················· | ···················· | ···················· | ···················· | |
| C | 288 hCoV-19/England/NORW-E8842/2020\|EPI_ISL_448240\|202 | ···················· | ···················· | ···················· | ···················· | ···················· | ···················· | |
| C | 289 hCoV-19/England/NORW-E8851/2020\|EPI_ISL_448241\|202 | ···················· | ···················· | ···················· | ···················· | ···················· | ···················· | |
| C | 290 hCoV-19/England/NORW-E888E/2020\|EPI_ISL_448242\|202 | ···················· | ···················· | ···················· | ···················· | ···················· | ···················· | |
| C | 291 hCoV-19/England/NORW-E889D/2020\|EPI_ISL_448243\|202 | ···················· | ···················· | ···················· | ···················· | ···················· | ···················· | |
| C | 292 hCoV-19/England/NORW-E88AC/2020\|EPI_ISL_448244\|202 | ···················· | ···················· | ···················· | ···················· | ···················· | ···················· | |
| C | 293 hCoV-19/England/NORW-E88BB/2020\|EPI_ISL_448245\|202 | ···················· | ···················· | ···················· | ···················· | ···················· | ···················· | |
| C | 294 hCoV-19/England/NORW-E88CA/2020\|EPI_ISL_448246\|202 | ···················· | ···················· | ···················· | ···················· | ···················· | ···················· | |
| C | 295 hCoV-19/England/NORW-E88D9/2020\|EPI_ISL_448247\|202 | ···················· | ···················· | ···················· | ···················· | ···················· | ···················· | |
| C | 296 hCoV-19/England/NORW-E88E8/2020\|EPI_ISL_448248\|202 | ···················· | ···················· | ···················· | ···················· | ···················· | ···················· | |
| C | 297 hCoV-19/England/NORW-E88F7/2020\|EPI_ISL_448249\|202 | ···················· | ···················· | ···················· | ···················· | ···················· | ···················· | |
| C | 298 hCoV-19/England/NORW-E8921/2020\|EPI_ISL_448250\|202 | ···················· | ···················· | ···················· | ···················· | ···················· | ···················· | |
| C | 299 hCoV-19/England/NORW-E8930/2020\|EPI_ISL_448251\|202 | ···················· | ···················· | ···················· | ···················· | ···················· | ···················· | |
| C | 300 hCoV-19/England/NORW-E895E/2020\|EPI_ISL_448253\|202 | ···················· | ···················· | ···················· | ···················· | ···················· | ···················· | |
| C | 301 hCoV-19/England/NORW-E896D/2020\|EPI_ISL_448254\|202 | ···················· | ···················· | ···················· | ···················· | ···················· | ···················· | |
| C | 302 hCoV-19/England/NORW-E897C/2020\|EPI_ISL_448255\|202 | ···················· | ···················· | ···················· | ···················· | ···················· | ···················· | |
| C | 303 hCoV-19/England/NORW-E898B/2020\|EPI_ISL_448256\|202 | ···················· | ···················· | ···················· | ···················· | ···················· | ···················· | |
| C | 305 hCoV-19/England/NORW-E89C7/2020\|EPI_ISL_448259\|202 | ···················· | ···················· | ···················· | ···················· | ···················· | ···················· | |
| C | 306 hCoV-19/Lebanon/S1_758/2020\|EPI_ISL_450508\|2020-03 | ···················· | ···················· | ···················· | ···················· | ···················· | ···················· | |
| C | 307 hCoV-19/Lebanon/S2_759/2020\|EPI_ISL_450509\|2020-03 | ···················· | ···················· | ···················· | ···················· | ···················· | ···················· | |
| C | 308 hCoV-19/Lebanon/S4_761/2020\|EPI_ISL_450511\|2020-03 | ···················· | ···················· | ···················· | ···················· | ···················· | ···················· | |
| C | 309 hCoV-19/Lebanon/S5_762/2020\|EPI_ISL_450512\|2020-02 | ···················· | ···················· | ···················· | ···················· | ···················· | ···················· | |
| C | 310 hCoV-19/Lebanon/S9_764/2020\|EPI_ISL_450515\|2020-03 | ···················· | ···················· | ···················· | ···················· | ···················· | ···················· | |
| C | 311 hCoV-19/Latvia/018/2020\|EPI_ISL_450518\|2020-04-18 | ···················· | ···················· | ···················· | ···················· | ···················· | ···················· | |
| C | 312 hCoV-19/Latvia/019/2020\|EPI_ISL_450519\|2020-04-14 | ···················· | ···················· | ···················· | ···················· | ···················· | ···················· | |
| C | 313 hCoV-19/Poland/1109500/2020\|EPI_ISL_450526\|2020-03 | ···················· | ···················· | ···················· | ···················· | ···················· | ···················· | |
| C | 314 hCoV-19/Poland/1110551/2020\|EPI_ISL_450527\|2020-03 | ···················· | ···················· | ···················· | ···················· | ···················· | ···················· | |
| C | 315 hCoV-19/Poland/1110540/2020\|EPI_ISL_450528\|2020-03 | ···················· | ···················· | ···················· | ···················· | ···················· | ···················· | |
| C | 316 hCoV-19/Poland/1111112/2020\|EPI_ISL_450529\|2020-03 | ···················· | ···················· | ···················· | ···················· | ···················· | ···················· | |
| C | 317 hCoV-19/Poland/1111628/2020\|EPI_ISL_450530\|2020-04 | ···················· | ···················· | ···················· | ···················· | ···················· | ···················· | |
| C | 318 hCoV-19/USA/UT-03794/2020\|EPI_ISL_450538\|2020-03-1 | ···················· | ···················· | ···················· | ···················· | ···················· | ···················· | |
| C | 319 hCoV-19/USA/UT-03470/2020\|EPI_ISL_450539\|2020-03-1 | ···················· | ···················· | ···················· | ···················· | ···················· | ···················· | |
| C | 320 hCoV-19/USA/UT-03474/2020\|EPI_ISL_450541\|2020-03-1 | ···················· | ···················· | ···················· | ···················· | ···················· | ···················· | |
| C | 321 hCoV-19/USA/UT-03475/2020\|EPI_ISL_450542\|2020-03-1 | ···················· | ···················· | ···················· | ···················· | ···················· | ···················· | |
| C | 322 hCoV-19/USA/UT-03477/2020\|EPI_ISL_450543\|2020-03-3 | ···················· | ···················· | ···················· | ···················· | ···················· | ···················· | |
| C | 323 hCoV-19/USA/UT-03482/2020\|EPI_ISL_450544\|2020-04-0 | ···················· | ···················· | ···················· | ···················· | ···················· | ···················· | |
| C | 324 hCoV-19/USA/UT-03484/2020\|EPI_ISL_450546\|2020-04-0 | ···················· | ···················· | ···················· | ···················· | ···················· | ···················· | |
| C | 325 hCoV-19/USA/UT-03485/2020\|EPI_ISL_450547\|2020-04-0 | ···················· | ···················· | ···················· | ···················· | ···················· | ···················· | |
| C | 326 hCoV-19/USA/UT-03486/2020\|EPI_ISL_450548\|2020-04-0 | ···················· | ···················· | ···················· | ···················· | ···················· | ···················· | |
| C | 327 hCoV-19/USA/UT-03696/2020\|EPI_ISL_450551\|2020-04-0 | ···················· | ···················· | ···················· | ···················· | ···················· | ···················· | |
| C | 328 hCoV-19/USA/UT-03698/2020\|EPI_ISL_450553\|2020-04-0 | ···················· | ···················· | ···················· | ···················· | ···················· | ···················· | |
| C | 329 hCoV-19/USA/UT-03699/2020\|EPI_ISL_450554\|2020-04-0 | ···················· | ···················· | ···················· | ···················· | ···················· | ···················· | |
| C | 330 hCoV-19/USA/UT-01234/2020\|EPI_ISL_450559\|2020-04-0 | ···················· | ···················· | ···················· | ···················· | ···················· | ···················· | |
| C | 331 hCoV-19/USA/UT-01236/2020\|EPI_ISL_450560\|2020-04-0 | ···················· | ···················· | ···················· | ···················· | ···················· | ···················· | |
| C | 332 hCoV-19/USA/UT-01237/2020\|EPI_ISL_450561\|2020-04-0 | ···················· | ···················· | ···················· | ···················· | ···················· | ···················· | |
| C | 333 hCoV-19/USA/UT-03878/2020\|EPI_ISL_450563\|2020-04-1 | ···················· | ···················· | ···················· | ···················· | ···················· | ···················· | |
| C | 334 hCoV-19/USA/UT-03879/2020\|EPI_ISL_450564\|2020-04-1 | ···················· | ···················· | ···················· | ···················· | ···················· | ···················· | |
| C | 335 hCoV-19/USA/UT-03889/2020\|EPI_ISL_450566\|2020-04-1 | ···················· | ···················· | ···················· | ···················· | ···················· | ···················· | |
| C | 336 hCoV-19/USA/UT-03909/2020\|EPI_ISL_450567\|2020-04-1 | ···················· | ···················· | ···················· | ···················· | ···················· | ···················· | |
| C | 337 hCoV-19/USA/MI-MDHHS-SC20361/2020\|EPI_ISL_450572\|2 | ···················· | ···················· | ···················· | ···················· | ···················· | ···················· | |
| C | 338 hCoV-19/Switzerland/GE-ETHZ-101135/2020\|EPI_ISL_45 | ···················· | ···················· | ···················· | ···················· | ·······S············ | ···················· | |
| C | 339 hCoV-19/Switzerland/BS-ETHZ-101136/2020\|EPI_ISL_45 | ···················· | ···················· | ···················· | ···················· | ·······S············ | ···················· | |
| C | 341 hCoV-19/Switzerland/BL-ETHZ-101143/2020\|EPI_ISL_45 | ···················· | ···················· | ···················· | ···················· | ···················· | ···················· | |
| C | 342 hCoV-19/Switzerland/BE-ETHZ-101144/2020\|EPI_ISL_45 | ···················· | ·A·················· | ···················· | ···················· | ···················· | ···················· | |

| | | | | | | | | |
|---|---|---|---|---|---|---|---|---|
| C | 343 | hCoV-19/Switzerland/BE-ETHZ-101145/2020\|EPI_ISL_45 | ···················· | ···················· | ···················· | ···················· | ···················· | ···················· |
| C | 344 | hCoV-19/Switzerland/BL-ETHZ-101146/2020\|EPI_ISL_45 | ···················· | ···················· | ···················· | ···················· | ···················· | ···················· |
| C | 348 | hCoV-19/Switzerland/BL-ETHZ-101150/2020\|EPI_ISL_45 | ···················· | ···················· | ···················· | ···················· | ···················· | ···················· |
| C | 350 | hCoV-19/Switzerland/BL-ETHZ-101152/2020\|EPI_ISL_45 | ···················· | ···················· | ···················· | ···················· | ···················· | ···················· |
| C | 351 | hCoV-19/Switzerland/BL-ETHZ-101155/2020\|EPI_ISL_45 | ···················· | ···················· | ···················· | ···················· | ···················· | ···················· |
| C | 352 | hCoV-19/Switzerland/BL-ETHZ-101156/2020\|EPI_ISL_45 | ···················· | ···················· | ···················· | ···················· | ·······S············ | ···················· |
| C | 354 | hCoV-19/Switzerland/AG-ETHZ-101158/2020\|EPI_ISL_45 | ···················· | ···················· | ···················· | ···················· | ···················· | ···················· |
| C | 355 | hCoV-19/Switzerland/BE-ETHZ-101160/2020\|EPI_ISL_45 | ···················· | ···················· | ···················· | ···················· | ·······S············ | ···················· |
| C | 357 | hCoV-19/Switzerland/AG-ETHZ-101165/2020\|EPI_ISL_45 | ···················· | ···················· | ···················· | ···················· | ···················· | ···················· |
| C | 360 | hCoV-19/Switzerland/GR-ETHZ-101168/2020\|EPI_ISL_45 | ···················· | ···················· | ···················· | ···················· | ···················· | ···················· |
| C | 361 | hCoV-19/Switzerland/GR-ETHZ-101170/2020\|EPI_ISL_45 | ···················· | ···················· | ···················· | ···················· | ···················· | ···················· |
| C | 362 | hCoV-19/Switzerland/ZH-ETHZ-101171/2020\|EPI_ISL_45 | ···················· | ···················· | ···················· | ···················· | ···················· | ···················· |
| C | 363 | hCoV-19/Switzerland/ZH-ETHZ-101172/2020\|EPI_ISL_45 | ···················· | ···················· | ···················· | ···················· | ·······S············ | ··············M····· |
| C | 364 | hCoV-19/Switzerland/ZH-ETHZ-101174/2020\|EPI_ISL_45 | ···················· | ···················· | ···················· | ···················· | ·················Y·· | ···················· |
| C | 366 | hCoV-19/Switzerland/GE-ETHZ-101177/2020\|EPI_ISL_45 | ···················· | ···················· | ···················· | ···················· | ···················· | ···················· |
| C | 367 | hCoV-19/Switzerland/ZH-ETHZ-101178/2020\|EPI_ISL_45 | ···················· | ···················· | ···················· | ···················· | ···················· | ···················· |
| C | 369 | hCoV-19/Switzerland/ZH-ETHZ-101181/2020\|EPI_ISL_45 | ···················· | ···················· | ···················· | ···················· | ···················· | ···················· |
| C | 370 | hCoV-19/Switzerland/BL-ETHZ-101182/2020\|EPI_ISL_45 | ···················· | ···················· | ···················· | ···················· | ·······S············ | ···················· |
| C | 371 | hCoV-19/Switzerland/BL-ETHZ-101184/2020\|EPI_ISL_45 | ···················· | ···················· | ···················· | ···················· | ···················· | ···················· |
| C | 372 | hCoV-19/Switzerland/SH-ETHZ-101186/2020\|EPI_ISL_45 | ···················· | ···················· | ···················· | ···················· | ···················· | ···················· |
| C | 376 | hCoV-19/Switzerland/ZH-ETHZ-101190/2020\|EPI_ISL_45 | ···················· | ···················· | ···················· | ···················· | ·······M············ | ···················· |
| C | 377 | hCoV-19/Switzerland/ZH-ETHZ-101191/2020\|EPI_ISL_45 | ···················· | ···················· | ···················· | ···················· | ···················· | ···················· |
| C | 378 | hCoV-19/Switzerland/AG-ETHZ-101192/2020\|EPI_ISL_45 | ···················· | ···················· | ···················· | ···················· | ···················· | ···················· |
| C | 379 | hCoV-19/Switzerland/ZH-ETHZ-101193/2020\|EPI_ISL_45 | ···················· | ···················· | ···················· | ···················· | ···················· | ···················· |
| C | 390 | hCoV-19/Switzerland/GL-ETHZ-110455/2020\|EPI_ISL_45 | ···················· | ···················· | ···················· | ···················· | ···················· | ···················· |
| C | 399 | hCoV-19/Switzerland/VD-ETHZ-110470/2020\|EPI_ISL_45 | ···················· | ·A·················· | ···················· | ···················· | ···················· | ···················· |
| C | 404 | hCoV-19/Portugal/PT0301/2020\|EPI_ISL_454017\|2020-0 | ···················· | ···················· | ···················· | ···················· | ···················· | ···················· |
| C | 405 | hCoV-19/Portugal/PT0302/2020\|EPI_ISL_454018\|2020-0 | ···················· | ···················· | ···················· | ···················· | ···················· | ···················· |
| C | 406 | hCoV-19/Portugal/PT0303/2020\|EPI_ISL_454019\|2020-0 | ···················· | ···················· | ···················· | ···················· | ···················· | ···················· |
| C | 407 | hCoV-19/Portugal/PT0304/2020\|EPI_ISL_454020\|2020-0 | ···················· | ···················· | ···················· | ···················· | ···················· | ···················· |
| C | 408 | hCoV-19/Portugal/PT0305/2020\|EPI_ISL_454021\|2020-0 | ···················· | ···················· | ···················· | ···················· | ···················· | ···················· |
| C | 409 | hCoV-19/Portugal/PT0306/2020\|EPI_ISL_454022\|2020-0 | ···················· | ···················· | ···················· | ···················· | ···················· | ···················· |
| C | 410 | hCoV-19/Portugal/PT0307/2020\|EPI_ISL_454023\|2020-0 | ···················· | ···················· | ···················· | ···················· | ···················· | ···················· |
| C | 411 | hCoV-19/Portugal/PT0309/2020\|EPI_ISL_454025\|2020-0 | ···················· | ···················· | ···················· | ···················· | ···················· | ···················· |
| C | 412 | hCoV-19/Portugal/PT0310/2020\|EPI_ISL_454026\|2020-0 | ···················· | ···················· | ···················· | ···················· | ···················· | ···················· |
| C | 413 | hCoV-19/Portugal/PT0313/2020\|EPI_ISL_454029\|2020-0 | ···················· | ···················· | ···················· | ···················· | ···················· | ···················· |
| C | 414 | hCoV-19/Portugal/PT0314/2020\|EPI_ISL_454030\|2020-0 | ···················· | ···················· | ···················· | ···················· | ···················· | ···················· |
| C | 415 | hCoV-19/Portugal/PT0315/2020\|EPI_ISL_454031\|2020-0 | ···················· | ···················· | ···················· | ···················· | ···················· | ···················· |
| C | 416 | hCoV-19/Portugal/PT0316/2020\|EPI_ISL_454032\|2020-0 | ···················· | ···················· | ···················· | ···················· | ···················· | ···················· |
| C | 417 | hCoV-19/Portugal/PT0317/2020\|EPI_ISL_454033\|2020-0 | ···················· | ···················· | ···················· | ···················· | ···················· | ···················· |
| C | 418 | hCoV-19/Portugal/PT0318/2020\|EPI_ISL_454034\|2020-0 | ···················· | ···················· | ···················· | ···················· | ···················· | ···················· |
| C | 419 | hCoV-19/Portugal/PT0319/2020\|EPI_ISL_454035\|2020-0 | ···················· | ···················· | ···················· | ···················· | ···················· | ···················· |
| C | 420 | hCoV-19/Portugal/PT0320/2020\|EPI_ISL_454036\|2020-0 | ···················· | ···················· | ···················· | ···················· | ···················· | ···················· |
| C | 421 | hCoV-19/Portugal/PT0321/2020\|EPI_ISL_454037\|2020-0 | ···················· | ···················· | ···················· | ···················· | ···················· | ···················· |
| C | 422 | hCoV-19/Portugal/PT0322/2020\|EPI_ISL_454038\|2020-0 | ···················· | ···················· | ···················· | ···················· | ···················· | ···················· |
| C | 423 | hCoV-19/Portugal/PT0323/2020\|EPI_ISL_454039\|2020-0 | ···················· | ···················· | ···················· | ···················· | ···················· | ···················· |
| C | 424 | hCoV-19/Portugal/PT0324/2020\|EPI_ISL_454040\|2020-0 | ···················· | ···················· | ···················· | ···················· | ···················· | ···················· |
| C | 425 | hCoV-19/Portugal/PT0325/2020\|EPI_ISL_454041\|2020-0 | ···················· | ···················· | ···················· | ···················· | ···················· | ···················· |
| C | 426 | hCoV-19/Portugal/PT0326/2020\|EPI_ISL_454042\|2020-0 | ···················· | ···················· | ···················· | ···················· | ···················· | ···················· |
| C | 427 | hCoV-19/Portugal/PT0327/2020\|EPI_ISL_454043\|2020-0 | ···················· | ···················· | ···················· | ···················· | ···················· | ···················· |
| C | 428 | hCoV-19/Portugal/PT0328/2020\|EPI_ISL_454044\|2020-0 | ···················· | ···················· | ···················· | ···················· | ···················· | ···················· |
| C | 429 | hCoV-19/Portugal/PT0329/2020\|EPI_ISL_454045\|2020-0 | ···················· | ···················· | ···················· | ···················· | ···················· | ···················· |
| C | 430 | hCoV-19/Portugal/PT0330/2020\|EPI_ISL_454046\|2020-0 | ···················· | ···················· | ···················· | ···················· | ···················· | ···················· |
| C | 431 | hCoV-19/Portugal/PT0331/2020\|EPI_ISL_454047\|2020-0 | ···················· | ···················· | ···················· | ···················· | ···················· | ···················· |
| C | 432 | hCoV-19/Portugal/PT0332/2020\|EPI_ISL_454048\|2020-0 | ···················· | ···················· | ···················· | ···················· | ···················· | ···················· |
| C | 433 | hCoV-19/Portugal/PT0333/2020\|EPI_ISL_454049\|2020-0 | ···················· | ···················· | ···················· | ···················· | ···················· | ···················· |
| C | 434 | hCoV-19/Portugal/PT0334/2020\|EPI_ISL_454050\|2020-0 | ···················· | ···················· | ···················· | ···················· | ···················· | ···················· |
| C | 435 | hCoV-19/Portugal/PT0335/2020\|EPI_ISL_454051\|2020-0 | ···················· | ···················· | ···················· | ···················· | ···················· | ···················· |
| C | 436 | hCoV-19/Portugal/PT0336/2020\|EPI_ISL_454052\|2020-0 | ···················· | ···················· | ···················· | ···················· | ···················· | ···················· |
| C | 437 | hCoV-19/Portugal/PT0337/2020\|EPI_ISL_454053\|2020-0 | ···················· | ···················· | ···················· | ···················· | ···················· | ···················· |
| C | 438 | hCoV-19/Portugal/PT0339/2020\|EPI_ISL_454055\|2020-0 | ···················· | ···················· | ···················· | ···················· | ···················· | ···················· |
| C | 439 | hCoV-19/Portugal/PT0341/2020\|EPI_ISL_454057\|2020-0 | ···················· | ···················· | ···················· | ···················· | ···················· | ···················· |
| C | 440 | hCoV-19/Portugal/PT0343/2020\|EPI_ISL_454059\|2020-0 | ···················· | ···················· | ···················· | ···················· | ···················· | ···················· |
| C | 441 | hCoV-19/Portugal/PT0374/2020\|EPI_ISL_454090\|2020-0 | ···················· | ···················· | ···················· | ···················· | ···················· | ···················· |
| C | 442 | hCoV-19/Portugal/PT0375/2020\|EPI_ISL_454091\|2020-0 | ···················· | ···················· | ···················· | ···················· | ···················· | ···················· |
| C | 443 | hCoV-19/Portugal/PT0376/2020\|EPI_ISL_454092\|2020-0 | ···················· | ···················· | ···················· | ···················· | ···················· | ···················· |
| C | 444 | hCoV-19/Portugal/PT0377/2020\|EPI_ISL_454093\|2020-0 | ···················· | ···················· | ···················· | ···················· | ···················· | ···················· |
| C | 445 | hCoV-19/Portugal/PT0378/2020\|EPI_ISL_454094\|2020-0 | ···················· | ···················· | ···················· | ···················· | ···················· | ···················· |
| C | 446 | hCoV-19/Portugal/PT0379/2020\|EPI_ISL_454095\|2020-0 | ···················· | ···················· | ···················· | ···················· | ···················· | ···················· |
| C | 447 | hCoV-19/Portugal/PT0380/2020\|EPI_ISL_454096\|2020-0 | ···················· | ···················· | ···················· | ···················· | ···················· | ···················· |
| C | 448 | hCoV-19/Portugal/PT0381/2020\|EPI_ISL_454097\|2020-0 | ···················· | ···················· | ···················· | ···················· | ···················· | ···················· |
| C | 449 | hCoV-19/Portugal/PT0382/2020\|EPI_ISL_454098\|2020-0 | ···················· | ···················· | ···················· | ···················· | ···················· | ···················· |
| C | 450 | hCoV-19/Portugal/PT0383/2020\|EPI_ISL_454099\|2020-0 | ···················· | ···················· | ···················· | ···················· | ···················· | ···················· |
| C | 451 | hCoV-19/Spain/ML-ISCIII-202449/2020\|EPI_ISL_455347 | ···················· | ···················· | ···················· | ···················· | ···················· | ···················· |
| C | 452 | hCoV-19/Spain/ML-ISCIII-202452/2020\|EPI_ISL_455348 | ···················· | ···················· | ···················· | ···················· | ···················· | ···················· |
| C | 453 | hCoV-19/Spain/ML-ISCIII-202454/2020\|EPI_ISL_455349 | ···················· | ···················· | ···················· | ···················· | ···················· | ···················· |
| C | 454 | hCoV-19/Spain/PV-ISCIII-201385/2020\|EPI_ISL_455350 | ···················· | ···················· | ···················· | ···················· | ···················· | ···················· |
| C | 455 | hCoV-19/Spain/PV-ISCIII-201388/2020\|EPI_ISL_455351 | ···················· | ···················· | ···················· | ···················· | ···················· | ···················· |

| | | | | | | | |
|---|---|---|---|---|---|---|---|
| C | 456 hCoV-19/Spain/PV-ISCIII-201491/2020\|EPI_ISL_455352 | ···················· | ···················· | ····················· | ···················· | ···················· | ···················· |
| C | 457 hCoV-19/Spain/PV-ISCIII-201492/2020\|EPI_ISL_455353 | ···················· | ···················· | ····················· | ···················· | ···················· | ···················· |
| C | 458 hCoV-19/Spain/PV-ISCIII-201920/2020\|EPI_ISL_455354 | ···················· | ···················· | ····················· | ···················· | ···················· | ···················· |
| C | 459 hCoV-19/USA/GA-EHC-009I/2020\|EPI_ISL_455355\|2020-0 | ···················· | ···················· | ····················· | ···················· | ···················· | ···················· |
| C | 460 hCoV-19/USA/GA-EHC-086H/2020\|EPI_ISL_455356\|2020-0 | ···················· | ···················· | ····················· | ···················· | ···················· | ···················· |
| C | 461 hCoV-19/USA/GA-EHC-136H/2020\|EPI_ISL_455358\|2020-0 | ···················· | ···················· | ····················· | ···················· | ···················· | ···················· |
| C | 462 hCoV-19/USA/GA-EHC-209C/2020\|EPI_ISL_455359\|2020-0 | ···················· | ···················· | ····················· | ···················· | ···················· | ···················· |
| C | 463 hCoV-19/USA/GA-EHC-087I/2020\|EPI_ISL_455360\|2020-0 | ···················· | ···················· | ····················· | ···················· | ···················· | ···················· |
| C | 464 hCoV-19/USA/GA-EHC-115L/2020\|EPI_ISL_455361\|2020-0 | ···················· | ···················· | ····················· | ···················· | ···················· | ···················· |
| C | 465 hCoV-19/Nigeria/KW017-CV24/2020\|EPI_ISL_455362\|202 | ···················· | ···················· | ····················· | ···················· | ···················· | ···················· |
| C | 466 hCoV-19/Wuhan/HB-WH1-123/2020\|EPI_ISL_455363\|2020- | ···················· | ···················· | ····················· | ···················· | ···················· | ···················· |
| C | 467 hCoV-19/Wuhan/HB-WH1-129/2020\|EPI_ISL_455364\|2020- | ···················· | ···················· | ····················· | ···················· | ···················· | ···················· |
| C | 468 hCoV-19/Wuhan/HB-WH1-132/2020\|EPI_ISL_455365\|2020- | ···················· | ···················· | ····················· | ···················· | ···················· | ···················· |
| C | 469 hCoV-19/Wuhan/HB-WH1-133/2020\|EPI_ISL_455366\|2020- | ···················· | ···················· | ····················· | ···················· | ···················· | ···················· |
| C | 470 hCoV-19/Wuhan/HB-WH1-139/2020\|EPI_ISL_455367\|2020- | ···················· | ···················· | ····················· | ···················· | ···················· | ···················· |
| C | 471 hCoV-19/Wuhan/HB-WH1-144/2020\|EPI_ISL_455368\|2020- | ···················· | ···················· | ····················· | ···················· | ···················· | ···················· |
| C | 472 hCoV-19/Wuhan/HB-WH2-145/2020\|EPI_ISL_455369\|2020- | ···················· | ···················· | ····················· | ···················· | ···················· | ···················· |
| C | 473 hCoV-19/Wuhan/HB-WH2-146/2020\|EPI_ISL_455370\|2020- | ···················· | ···················· | ····················· | ···················· | ···················· | ···················· |
| C | 474 hCoV-19/Wuhan/HB-WH2-147/2020\|EPI_ISL_455371\|2020- | ···················· | ···················· | ····················· | ···················· | ···················· | ···················· |
| C | 475 hCoV-19/Wuhan/HB-WH2-151/2020\|EPI_ISL_455372\|2020- | ···················· | ···················· | ····················· | ···················· | ···················· | ···················· |
| C | 476 hCoV-19/Wuhan/HB-WH2-154/2020\|EPI_ISL_455373\|2020- | ···················· | ···················· | ····················· | ···················· | ···················· | ···················· |
| C | 477 hCoV-19/Wuhan/HB-WH2-162/2020\|EPI_ISL_455374\|2020- | ···················· | ···················· | ····················· | ···················· | ···················· | ···················· |
| C | 478 hCoV-19/Wuhan/HB-WH2-165/2020\|EPI_ISL_455375\|2020- | ···················· | ···················· | ····················· | ···················· | ···················· | ···················· |
| C | 479 hCoV-19/Wuhan/HB-WH3-174/2020\|EPI_ISL_455376\|2020- | ···················· | ···················· | ····················· | ···················· | ···················· | ···················· |
| C | 480 hCoV-19/Wuhan/HB-WH3-177/2020\|EPI_ISL_455377\|2020- | ···················· | ···················· | ····················· | ···················· | ···················· | ···················· |
| C | 481 hCoV-19/Wuhan/HB-WH3-178/2020\|EPI_ISL_455378\|2020- | ···················· | ···················· | ····················· | ···················· | ···················· | ···················· |
| C | 482 hCoV-19/Wuhan/HB-WH3-180/2020\|EPI_ISL_455379\|2020- | ···················· | ···················· | ····················· | ···················· | ···················· | ···················· |
| C | 483 hCoV-19/Wuhan/HB-WH3-184/2020\|EPI_ISL_455380\|2020- | ···················· | ···················· | ····················· | ···················· | ···················· | ···················· |
| C | 484 hCoV-19/Wuhan/HB-WH3-186/2020\|EPI_ISL_455381\|2020- | ···················· | ···················· | ····················· | ···················· | ···················· | ···················· |
| C | 485 hCoV-19/Wuhan/HB-WH3-187/2020\|EPI_ISL_455382\|2020- | ···················· | ···················· | ····················· | ···················· | ···················· | ···················· |
| C | 486 hCoV-19/Wuhan/HB-WH3-188/2020\|EPI_ISL_455383\|2020- | ···················· | ···················· | ····················· | ···················· | ···················· | ···················· |
| C | 487 hCoV-19/Wuhan/HB-WH3-190/2020\|EPI_ISL_455384\|2020- | ···················· | ···················· | ····················· | ···················· | ···················· | ···················· |
| C | 488 hCoV-19/Wuhan/HB-WH4-198/2020\|EPI_ISL_455386\|2020- | ···················· | ···················· | ····················· | ···················· | ···················· | ···················· |
| C | 489 hCoV-19/Wuhan/HB-WH4-202/2020\|EPI_ISL_455388\|2020- | ···················· | ···················· | ····················· | ···················· | ···················· | ···················· |
| C | 490 hCoV-19/Wuhan/HB-WH4-207/2020\|EPI_ISL_455389\|2020- | ···················· | ···················· | ····················· | ···················· | ···················· | ···················· |
| C | 491 hCoV-19/England/CAMB-1B245B/2020\|EPI_ISL_456680\|20 | ···················· | ···················· | ····················· | ···················· | ···················· | ···················· |
| C | 494 hCoV-19/England/CAMB-1B25FE/2020\|EPI_ISL_456699\|20 | ···················· | ···················· | ····················· | ···················· | ···················· | ···················· |
| C | 495 hCoV-19/Netherlands/NoordBrabant_166/2020\|EPI_ISL_ | ···················· | ···················· | ····················· | ···················· | ···················· | ···················· |
| C | 496 hCoV-19/Netherlands/NoordBrabant_167/2020\|EPI_ISL_ | ···················· | ···················· | ····················· | ···················· | ···················· | ···················· |
| C | 497 hCoV-19/Netherlands/NoordBrabant_168/2020\|EPI_ISL_ | ···················· | ···················· | ····················· | ···················· | ···················· | ···················· |
| C | 498 hCoV-19/Netherlands/NoordHolland_14/2020\|EPI_ISL_4 | ···················· | ···················· | ····················· | ···················· | ···················· | ···················· |
| C | 499 hCoV-19/Netherlands/NoordHolland_16/2020\|EPI_ISL_4 | ···················· | ···················· | ····················· | ···················· | ···················· | ···················· |
| C | 500 hCoV-19/Netherlands/NoordHolland_17/2020\|EPI_ISL_4 | ···················· | ···················· | ····················· | ···················· | ···················· | ···················· |
| C | 501 hCoV-19/Netherlands/NoordHolland_18/2020\|EPI_ISL_4 | ···················· | ···················· | ····················· | ···················· | ···················· | ···················· |
| C | 502 hCoV-19/Netherlands/NoordHolland_19/2020\|EPI_ISL_4 | ···················· | ···················· | ····················· | ···················· | ···················· | ···················· |
| C | 503 hCoV-19/Netherlands/NoordHolland_20/2020\|EPI_ISL_4 | ···················· | ···················· | ····················· | ···················· | ···················· | ···················· |
| C | 504 hCoV-19/Netherlands/NoordHolland_21/2020\|EPI_ISL_4 | ···················· | ···················· | ····················· | ···················· | ···················· | ···················· |
| C | 505 hCoV-19/Netherlands/NoordHolland_22/2020\|EPI_ISL_4 | ···················· | ···················· | ····················· | ···················· | ···················· | ···················· |
| C | 506 hCoV-19/Netherlands/NoordHolland_23/2020\|EPI_ISL_4 | ···················· | ···················· | ····················· | ···················· | ···················· | ···················· |
| C | 507 hCoV-19/Netherlands/NoordHolland_24/2020\|EPI_ISL_4 | ···················· | ···················· | ····················· | ···················· | ···················· | ···················· |
| C | 508 hCoV-19/Netherlands/NoordHolland_25/2020\|EPI_ISL_4 | ···················· | ···················· | ····················· | ···················· | ···················· | ···················· |
| C | 509 hCoV-19/Netherlands/NoordHolland_26/2020\|EPI_ISL_4 | ···················· | ···················· | ····················· | ···················· | ···················· | ···················· |
| C | 510 hCoV-19/Netherlands/NoordHolland_27/2020\|EPI_ISL_4 | ···················· | ···················· | ····················· | ···················· | ···················· | ···················· |
| C | 511 hCoV-19/Netherlands/NoordHolland_28/2020\|EPI_ISL_4 | ···················· | ···················· | ····················· | ···················· | ···················· | ···················· |
| C | 512 hCoV-19/Netherlands/NoordHolland_29/2020\|EPI_ISL_4 | ···················· | ···················· | ····················· | ···················· | ···················· | ···················· |
| C | 513 hCoV-19/England/NORW-E9EF4/2020\|EPI_ISL_448380\|202 | ···················· | ···················· | ···········T········· | ···················· | ···················· | ···················· |
| C | 514 hCoV-19/England/NORW-E9FE2/2020\|EPI_ISL_448383\|202 | ···················· | ···················· | ····················· | ···················· | ···················· | ···················· |
| C | 515 hCoV-19/England/NORW-E9FF1/2020\|EPI_ISL_448384\|202 | ···················· | ···················· | ····················· | ···················· | ···················· | ···················· |
| C | 516 hCoV-19/England/NORW-EA00C/2020\|EPI_ISL_448385\|202 | ···················· | ···················· | ····················· | ···················· | ···················· | ···················· |
| C | 517 hCoV-19/England/NORW-EA057/2020\|EPI_ISL_448387\|202 | ···················· | ···················· | ···········T········· | ···················· | ···················· | ···················· |
| C | 518 hCoV-19/England/NORW-EA109/2020\|EPI_ISL_448388\|202 | ···················· | ···················· | ····················· | ···················· | ···················· | ···················· |
| C | 519 hCoV-19/England/NOTT-110AFF/2020\|EPI_ISL_448389\|20 | ···················· | ···················· | ····················· | ···················· | ···················· | ···················· |
| C | 520 hCoV-19/England/NOTT-110B74/2020\|EPI_ISL_448391\|20 | ···················· | ···················· | ····················· | ···················· | ···················· | ···················· |
| C | 521 hCoV-19/England/NOTT-110C08/2020\|EPI_ISL_448393\|20 | ···················· | ···················· | ····················· | ···················· | ···················· | ···················· |
| C | 522 hCoV-19/England/NOTT-110C26/2020\|EPI_ISL_448395\|20 | ···················· | ···················· | ····················· | ···················· | ···················· | ···················· |
| C | 523 hCoV-19/England/NOTT-110C44/2020\|EPI_ISL_448397\|20 | ···················· | ···················· | ····················· | ···················· | ···················· | ···················· |
| C | 524 hCoV-19/England/NOTT-110C62/2020\|EPI_ISL_448398\|20 | ···················· | ···················· | ····················· | ···················· | ···················· | ···················· |
| C | 525 hCoV-19/England/NOTT-110C71/2020\|EPI_ISL_448399\|20 | ···················· | ···················· | ····················· | ···················· | ···················· | ···················· |
| C | 526 hCoV-19/Switzerland/ZH-ETHZ-101133/2020\|EPI_ISL_45 | ···················· | ···················· | ····················· | ···················· | ···················· | ···················· |
| C | 528 hCoV-19/Switzerland/ZH-ETHZ-101138/2020\|EPI_ISL_45 | ···················· | ···················· | ····················· | ···················· | ···················· | ···················· |
| C | 529 hCoV-19/Switzerland/BE-ETHZ-101139/2020\|EPI_ISL_45 | ···················· | ·A·················· | ····················· | ···················· | ··········M········· | ···················· |
| C | 530 hCoV-19/Switzerland/BL-ETHZ-101140/2020\|EPI_ISL_45 | ···················· | ···················· | ····················· | ···················· | ···················· | ···················· |
| C | 532 hCoV-19/Switzerland/BL-ETHZ-101142/2020\|EPI_ISL_45 | ···················· | ···················· | ····················· | ···················· | ·······S············ | ···················· |
| C | 533 hCoV-19/Portugal/PT0344/2020\|EPI_ISL_454060\|2020-0 | ···················· | ···················· | ····················· | ···················· | ···················· | ···················· |
| C | 534 hCoV-19/Portugal/PT0345/2020\|EPI_ISL_454061\|2020-0 | ···················· | ···················· | ····················· | ···················· | ···················· | ···················· |
| C | 535 hCoV-19/Portugal/PT0346/2020\|EPI_ISL_454062\|2020-0 | ···················· | ···················· | ····················· | ···················· | ···················· | ···················· |
| C | 536 hCoV-19/Portugal/PT0347/2020\|EPI_ISL_454063\|2020-0 | ···················· | ···················· | ····················· | ···················· | ···················· | ···················· |
| C | 537 hCoV-19/Portugal/PT0348/2020\|EPI_ISL_454064\|2020-0 | ···················· | ···················· | ····················· | ···················· | ···················· | ···················· |

| | | | | | | | | |
|---|---|---|---|---|---|---|---|---|
| C | 538 | hCoV-19/Portugal/PT0349/2020\|EPI_ISL_454065\|2020-0 | .................... | .................... | ..................... | .................... | .................... | .................... |
| C | 539 | hCoV-19/Portugal/PT0350/2020\|EPI_ISL_454066\|2020-0 | .................... | .................... | ..................... | .................... | .................... | .................... |
| C | 540 | hCoV-19/Portugal/PT0351/2020\|EPI_ISL_454067\|2020-0 | .................... | .................... | ..................... | .................... | .................... | .................... |
| C | 541 | hCoV-19/Portugal/PT0352/2020\|EPI_ISL_454068\|2020-0 | .................... | .................... | ..................... | .................... | .................... | .................... |
| C | 542 | hCoV-19/Portugal/PT0353/2020\|EPI_ISL_454069\|2020-0 | .................... | .................... | ..................... | .................... | .................... | .................... |
| C | 543 | hCoV-19/Portugal/PT0354/2020\|EPI_ISL_454070\|2020-0 | .................... | .................... | ..................... | .................... | .................... | .................... |
| C | 544 | hCoV-19/Portugal/PT0355/2020\|EPI_ISL_454071\|2020-0 | .................... | .................... | ..................... | .................... | .................... | .................... |
| C | 545 | hCoV-19/Portugal/PT0356/2020\|EPI_ISL_454072\|2020-0 | .................... | .................... | ..................... | .................... | .................... | .................... |
| C | 546 | hCoV-19/Portugal/PT0358/2020\|EPI_ISL_454074\|2020-0 | .................... | .................... | ..................... | .................... | .................... | .................... |
| C | 547 | hCoV-19/Portugal/PT0360/2020\|EPI_ISL_454076\|2020-0 | .................... | .................... | ..................... | .................... | .................... | .................... |
| C | 548 | hCoV-19/Portugal/PT0361/2020\|EPI_ISL_454077\|2020-0 | .................... | .................... | ..................... | .................... | .................... | .................... |
| C | 549 | hCoV-19/Portugal/PT0362/2020\|EPI_ISL_454078\|2020-0 | .................... | .................... | ..................... | .................... | .................... | .................... |
| C | 550 | hCoV-19/Portugal/PT0364/2020\|EPI_ISL_454080\|2020-0 | .................... | .................... | ..................... | .................... | .................... | .................... |
| C | 551 | hCoV-19/Portugal/PT0365/2020\|EPI_ISL_454081\|2020-0 | .................... | .................... | ..................... | .................... | .................... | .................... |
| C | 552 | hCoV-19/Portugal/PT0366/2020\|EPI_ISL_454082\|2020-0 | .................... | .................... | ..................... | .................... | .................... | .................... |
| C | 553 | hCoV-19/Portugal/PT0367/2020\|EPI_ISL_454083\|2020-0 | .................... | .................... | ..................... | .................... | .................... | .................... |
| C | 554 | hCoV-19/Portugal/PT0369/2020\|EPI_ISL_454085\|2020-0 | .................... | .................... | ..................... | .................... | .................... | .................... |
| C | 555 | hCoV-19/Portugal/PT0370/2020\|EPI_ISL_454086\|2020-0 | .................... | .................... | ..................... | .................... | .................... | .................... |
| C | 556 | hCoV-19/Portugal/PT0371/2020\|EPI_ISL_454087\|2020-0 | .................... | .................... | ..................... | .................... | .................... | .................... |
| C | 557 | hCoV-19/Portugal/PT0373/2020\|EPI_ISL_454089\|2020-0 | .................... | .................... | ..................... | .................... | .................... | .................... |
| C | 558 | hCoV-19/Portugal/PT0476/2020\|EPI_ISL_454200\|2020-0 | .................... | .................... | ..................... | .................... | .................... | .................... |
| C | 559 | hCoV-19/Portugal/PT0477/2020\|EPI_ISL_454201\|2020-0 | .................... | .................... | ..................... | .................... | .................... | .................... |
| C | 560 | hCoV-19/Portugal/PT0478/2020\|EPI_ISL_454202\|2020-0 | .................... | .................... | ..................... | .................... | .................... | .................... |
| C | 561 | hCoV-19/Portugal/PT0479/2020\|EPI_ISL_454203\|2020-0 | .................... | .................... | ..................... | .................... | .................... | .................... |
| C | 562 | hCoV-19/Portugal/PT0480/2020\|EPI_ISL_454204\|2020-0 | .................... | .................... | ..................... | .................... | .................... | .................... |
| C | 563 | hCoV-19/Portugal/PT0481/2020\|EPI_ISL_454205\|2020-0 | .................... | .................... | ..................... | .................... | .................... | .................... |
| C | 564 | hCoV-19/Portugal/PT0482/2020\|EPI_ISL_454206\|2020-0 | .................... | .................... | ..................... | .................... | .................... | .................... |
| C | 565 | hCoV-19/Portugal/PT0483/2020\|EPI_ISL_454207\|2020-0 | .................... | .................... | ..................... | .................... | .................... | .................... |
| C | 566 | hCoV-19/Portugal/PT0484/2020\|EPI_ISL_454208\|2020-0 | .................... | .................... | ..................... | .................... | .................... | .................... |
| C | 567 | hCoV-19/Portugal/PT0485/2020\|EPI_ISL_454209\|2020-0 | .................... | .................... | ..................... | .................... | .................... | .................... |
| C | 568 | hCoV-19/Portugal/PT0486/2020\|EPI_ISL_454210\|2020-0 | .................... | .................... | ..................... | .................... | .................... | .................... |
| C | 569 | hCoV-19/Portugal/PT0487/2020\|EPI_ISL_454211\|2020-0 | .................... | .................... | ..................... | .................... | .................... | .................... |
| C | 570 | hCoV-19/Portugal/PT0489/2020\|EPI_ISL_454213\|2020-0 | .................... | .................... | ..................... | .................... | .................... | .................... |
| C | 571 | hCoV-19/Portugal/PT0490/2020\|EPI_ISL_454214\|2020-0 | .................... | .................... | ..................... | .................... | .................... | .................... |
| C | 572 | hCoV-19/Portugal/PT0491/2020\|EPI_ISL_454215\|2020-0 | .................... | .................... | ..................... | .................... | .................... | .................... |
| C | 573 | hCoV-19/Portugal/PT0492/2020\|EPI_ISL_454216\|2020-0 | .................... | .................... | ..................... | .................... | .................... | .................... |
| C | 574 | hCoV-19/Portugal/PT0493/2020\|EPI_ISL_454217\|2020-0 | .................... | .................... | ..................... | .................... | .................... | .................... |
| C | 575 | hCoV-19/Portugal/PT0494/2020\|EPI_ISL_454218\|2020-0 | .................... | .................... | ..................... | .................... | .................... | .................... |
| C | 576 | hCoV-19/Portugal/PT0495/2020\|EPI_ISL_454219\|2020-0 | .................... | .................... | ..................... | .................... | .................... | .................... |
| C | 577 | hCoV-19/Portugal/PT0496/2020\|EPI_ISL_454220\|2020-0 | .................... | .................... | ..................... | .................... | .................... | .................... |
| C | 578 | hCoV-19/Portugal/PT0497/2020\|EPI_ISL_454221\|2020-0 | .................... | .................... | ..................... | .................... | .................... | .................... |
| C | 579 | hCoV-19/Portugal/PT0498/2020\|EPI_ISL_454222\|2020-0 | .................... | .................... | ..................... | .................... | .................... | .................... |
| C | 580 | hCoV-19/Portugal/PT0499/2020\|EPI_ISL_454223\|2020-0 | .................... | .................... | ..................... | .................... | .................... | .................... |
| C | 581 | hCoV-19/Portugal/PT0500/2020\|EPI_ISL_454224\|2020-0 | .................... | .................... | ..................... | .................... | .................... | .................... |
| C | 582 | hCoV-19/Portugal/PT0501/2020\|EPI_ISL_454225\|2020-0 | .................... | .................... | ..................... | .................... | .................... | .................... |
| C | 583 | hCoV-19/Portugal/PT0502/2020\|EPI_ISL_454226\|2020-0 | .................... | .................... | ..................... | .................... | .................... | .................... |
| C | 584 | hCoV-19/Portugal/PT0503/2020\|EPI_ISL_454227\|2020-0 | .................... | .................... | ..................... | .................... | .................... | .................... |
| C | 585 | hCoV-19/Portugal/PT0504/2020\|EPI_ISL_454228\|2020-0 | .................... | .................... | ..................... | .................... | .................... | .................... |
| C | 586 | hCoV-19/Portugal/PT0505/2020\|EPI_ISL_454229\|2020-0 | .................... | .................... | ..................... | .................... | .................... | .................... |
| C | 587 | hCoV-19/Portugal/PT0506/2020\|EPI_ISL_454230\|2020-0 | .................... | .................... | ..................... | .................... | .................... | .................... |
| C | 588 | hCoV-19/Portugal/PT0508/2020\|EPI_ISL_454232\|2020-0 | .................... | .................... | ..................... | .................... | .................... | .................... |
| C | 589 | hCoV-19/Portugal/PT0509/2020\|EPI_ISL_454233\|2020-0 | .................... | .................... | ..................... | .................... | .................... | .................... |
| C | 590 | hCoV-19/Portugal/PT0510/2020\|EPI_ISL_454234\|2020-0 | .................... | .................... | ..................... | .................... | .................... | .................... |
| C | 591 | hCoV-19/Portugal/PT0511/2020\|EPI_ISL_454235\|2020-0 | .................... | .................... | ..................... | .................... | .................... | .................... |
| C | 592 | hCoV-19/Portugal/PT0512/2020\|EPI_ISL_454236\|2020-0 | .................... | .................... | ..................... | .................... | .................... | .................... |
| C | 593 | hCoV-19/Portugal/PT0513/2020\|EPI_ISL_454237\|2020-0 | .................... | .................... | ..................... | .................... | .................... | .................... |
| C | 594 | hCoV-19/Portugal/PT0514/2020\|EPI_ISL_454238\|2020-0 | .................... | .................... | ..................... | .................... | .................... | .................... |
| C | 595 | hCoV-19/Portugal/PT0516/2020\|EPI_ISL_454240\|2020-0 | .................... | .................... | ..................... | .................... | .................... | .................... |
| C | 596 | hCoV-19/Portugal/PT0517/2020\|EPI_ISL_454241\|2020-0 | .................... | .................... | ..................... | .................... | .................... | .................... |
| C | 597 | hCoV-19/Portugal/PT0518/2020\|EPI_ISL_454242\|2020-0 | .................... | .................... | ..................... | .................... | .................... | .................... |
| C | 598 | hCoV-19/Portugal/PT0519/2020\|EPI_ISL_454243\|2020-0 | .................... | .................... | ..................... | .................... | .................... | .................... |
| C | 599 | hCoV-19/Portugal/PT0520/2020\|EPI_ISL_454244\|2020-0 | .................... | .................... | ..................... | .................... | .................... | .................... |
| C | 600 | hCoV-19/Portugal/PT0521/2020\|EPI_ISL_454245\|2020-0 | .................... | .................... | ..................... | .................... | .................... | .................... |
| C | 601 | hCoV-19/Portugal/PT0522/2020\|EPI_ISL_454246\|2020-0 | .................... | .................... | ..................... | .................... | .................... | .................... |
| C | 602 | hCoV-19/Portugal/PT0523/2020\|EPI_ISL_454247\|2020-0 | .................... | .................... | ..................... | .................... | .................... | .................... |
| C | 603 | hCoV-19/Portugal/PT0524/2020\|EPI_ISL_454248\|2020-0 | .................... | .................... | ..................... | .................... | .................... | .................... |
| C | 604 | hCoV-19/Portugal/PT0525/2020\|EPI_ISL_454249\|2020-0 | .................... | .................... | ..................... | .................... | .................... | .................... |
| C | 605 | hCoV-19/Portugal/PT0526/2020\|EPI_ISL_454250\|2020-0 | .................... | .................... | ..................... | .................... | .................... | .................... |
| C | 606 | hCoV-19/Portugal/PT0527/2020\|EPI_ISL_454251\|2020-0 | .................... | .................... | ..................... | .................... | .................... | .................... |
| C | 607 | hCoV-19/Portugal/PT0528/2020\|EPI_ISL_454252\|2020-0 | .................... | .................... | ..................... | .................... | .................... | .................... |
| C | 608 | hCoV-19/Portugal/PT0529/2020\|EPI_ISL_454253\|2020-0 | .................... | .................... | ..................... | .................... | .................... | .................... |
| C | 609 | hCoV-19/Portugal/PT0531/2020\|EPI_ISL_454255\|2020-0 | .................... | .................... | ..................... | .................... | .................... | .................... |
| C | 610 | hCoV-19/Portugal/PT0533/2020\|EPI_ISL_454257\|2020-0 | .................... | .................... | ..................... | .................... | .................... | .................... |
| C | 611 | hCoV-19/Wuhan/HB-WH5-218/2020\|EPI_ISL_455390\|2020- | .................... | .................... | ..................... | .................... | .................... | .................... |
| C | 612 | hCoV-19/Wuhan/HB-WH5-228/2020\|EPI_ISL_455391\|2020- | .................... | .................... | ..................... | .................... | ...C................ | .................... |
| C | 613 | hCoV-19/Wuhan/HB-WH5-230/2020\|EPI_ISL_455392\|2020- | .................... | .................... | ..................... | .................... | .................... | .................... |
| C | 614 | hCoV-19/Wuhan/HB-WH5-235/2020\|EPI_ISL_455394\|2020- | .................... | .................... | ..................... | .................... | .................... | .................... |
| C | 615 | hCoV-19/Wuhan/HB-WH6-241/2020\|EPI_ISL_455395\|2020- | .................... | .................... | ..................... | .................... | .................... | .................... |

```
C    616 hCoV-19/Wuhan/HB-WH6-244/2020|EPI_ISL_455397|2020-   ····················  ····················  ····················  ····················  ····················  ····················
C    617 hCoV-19/Wuhan/HB-WH6-245/2020|EPI_ISL_455398|2020-   ····················  ····················  ····················  ····················  ····················  ····················
C    618 hCoV-19/Wuhan/HB-WHCM-101/2020|EPI_ISL_455399|2020   ····················  ····················  ····················  ····················  ····················  ····················
C    619 hCoV-19/Croatia/17-S7new/2020|EPI_ISL_455566|2020-   ····················  ····················  ····················  ····················  ····················  ····················
C    620 hCoV-19/Croatia/7R-S8new/2020|EPI_ISL_455567|2020-   ····················  ····················  ····················  ····················  ····················  ····················
C    621 hCoV-19/USA/IA-GMF-02699/2020|EPI_ISL_455568|2020-   ····················  ····················  ····················  ····················  ····················  ····················
C    622 hCoV-19/USA/WI-GMF-02728/2020|EPI_ISL_455569|2020-   ····················  ····················  ····················  ····················  ····················  ····················
C    623 hCoV-19/USA/MN-GMF-02870/2020|EPI_ISL_455570|2020-   ····················  ····················  ····················  ····················  ····················  ····················
C    624 hCoV-19/USA/WI-GMF-03151/2020|EPI_ISL_455571|2020-   ····················  ····················  ····················  ····················  ····················  ····················
C    625 hCoV-19/USA/WI-GMF-03156/2020|EPI_ISL_455572|2020-   ····················  ····················  ····················  ····················  ····················  ····················
C    626 hCoV-19/USA/WI-GMF-M00001/2020|EPI_ISL_455573|2020   ····················  ····················  ····················  ····················  ····················  ····················
C    627 hCoV-19/USA/WI-GMF-M00002/2020|EPI_ISL_455574|2020   ····················  ····················  ····················  ····················  ····················  ····················
C    628 hCoV-19/USA/WI-GMF-03710/2020|EPI_ISL_455575|2020-   ····················  ····················  ····················  ····················  ····················  ····················
C    629 hCoV-19/USA/WI-GMF-03823/2020|EPI_ISL_455576|2020-   ····················  ····················  ····················  ····················  ····················  ····················
C    630 hCoV-19/USA/WI-GMF-04314/2020|EPI_ISL_455578|2020-   ····················  ····················  ····················  ····················  ····················  ····················
C    631 hCoV-19/USA/WI-GMF-M00003/2020|EPI_ISL_455580|2020   ····················  ····················  ····················  ····················  ····················  ····················
C    632 hCoV-19/USA/WI-GMF-M00004/2020|EPI_ISL_455581|2020   ····················  ····················  ····················  ····················  ····················  ····················
C    633 hCoV-19/USA/IA-GMF-M00005/2020|EPI_ISL_455582|2020   ····················  ····················  ····················  ····················  ····················  ····················
C    634 hCoV-19/Thailand/Nonthaburi_2982/2020|EPI_ISL_4555   ····················  ····················  ····················  ····················  ····················  ····················
C    635 hCoV-19/Thailand/Phatum thani_2980/2020|EPI_ISL_45   ····················  ····················  ····················  ····················  ····················  ····················
C    636 hCoV-19/Thailand/Bangkok_2974/2020|EPI_ISL_455585|   ····················  ····················  ····················  ····················  ····················  ····················
C    637 hCoV-19/Thailand/Bangkok_2972/2020|EPI_ISL_455586|   ····················  ····················  ····················  ····················  ····················  ····················
C    638 hCoV-19/Thailand/Nakhonnayok_2829/2020|EPI_ISL_455   ····················  ····················  ····················  ····················  ····················  ····················
C    639 hCoV-19/Thailand/Trang_5008/2020|EPI_ISL_455588|20   ····················  ····················  ····················  ····················  ····················  ····················
C    641 hCoV-19/Scotland/CVR3410/2020|EPI_ISL_456834|2020-   ····················  ····················  ····················  ····················  ····················  ····················
C    642 hCoV-19/Scotland/CVR3411/2020|EPI_ISL_456835|2020-   ····················  ·A··················  ····················  ····················  ····················  ····················
C    644 hCoV-19/Scotland/CVR3416/2020|EPI_ISL_456839|2020-   ····················  ····················  ····················  ····················  ····················  ····················
C    652 hCoV-19/Scotland/CVR3445/2020|EPI_ISL_456855|2020-   ····················  ····················  ····················  ····················  ····················  ····················
C    654 hCoV-19/Scotland/CVR3469/2020|EPI_ISL_456863|2020-   ····················  ····················  ····················  ····················  ····················  ····················
C    655 hCoV-19/Scotland/CVR3470/2020|EPI_ISL_456864|2020-   ····················  ····················  ····················  ····················  ····················  ····················
C    656 hCoV-19/Scotland/CVR3471/2020|EPI_ISL_456865|2020-   ····················  ····················  ····················  ····················  ····················  ····················
C    657 hCoV-19/Scotland/CVR3472/2020|EPI_ISL_456866|2020-   ····················  ····················  ····················  ····················  ····················  ····················
C    658 hCoV-19/Scotland/CVR3474/2020|EPI_ISL_456868|2020-   ····················  ···A················  ····················  ····················  ····················  ····················
C    662 hCoV-19/Scotland/CVR3482/2020|EPI_ISL_456874|2020-   ····················  ····················  ····················  ····················  ····················  ····················
C    663 hCoV-19/Scotland/CVR3483/2020|EPI_ISL_456875|2020-   ····················  ····················  ····················  ····················  ····················  ····················
C    664 hCoV-19/Scotland/CVR3486/2020|EPI_ISL_456877|2020-   ····················  ·A··················  ····················  ····················  ····················  ····················
C    665 hCoV-19/Scotland/CVR3489/2020|EPI_ISL_456878|2020-   ····················  ····················  ····················  ····················  ····················  ····················
C    666 hCoV-19/Scotland/CVR3492/2020|EPI_ISL_456880|2020-   ····················  ····················  ····················  ····················  ····················  ····················
C    667 hCoV-19/Scotland/CVR3497/2020|EPI_ISL_456883|2020-   ····················  ····················  ····················  ····················  ····················  ····················
C    670 hCoV-19/Scotland/CVR3501/2020|EPI_ISL_456886|2020-   ····················  ····················  ····················  ····················  ····················  ····················
C    671 hCoV-19/Scotland/CVR3502/2020|EPI_ISL_456887|2020-   ····················  ····················  ····················  ····················  ····················  ····················
C    672 hCoV-19/Scotland/CVR853/2020|EPI_ISL_456889|2020-0   ····················  ····················  ····················  ····················  ····················  ····················
C    673 hCoV-19/Scotland/EDB1067/2020|EPI_ISL_456890|2020-   ····················  ····················  ····················  ····················  ····················  ····················
C    674 hCoV-19/Scotland/EDB1156/2020|EPI_ISL_456891|2020-   ····················  ····················  ····················  ····················  ····················  ····················
C    675 hCoV-19/Scotland/EDB1310/2020|EPI_ISL_456893|2020-   ····················  ····················  ····················  ····················  ····················  ····················
C    676 hCoV-19/Scotland/EDB213/2020|EPI_ISL_456894|2020-0   ····················  ····················  ····················  ····················  ····················  ····················
C    677 hCoV-19/Scotland/EDB2730/2020|EPI_ISL_456896|2020-   ····················  ···················A  ····················  ····················  ····················  ····················     1
C    678 hCoV-19/Scotland/EDB2742/2020|EPI_ISL_456897|2020-   ····················  ····················  ····················  ····················  ····················  ····················
C    679 hCoV-19/Scotland/EDB2778/2020|EPI_ISL_456898|2020-   ····················  ····················  ····················  ····················  ····················  ····················
C    680 hCoV-19/England/BRIS-125FF8/2020|EPI_ISL_459035|20   ····················  ····················  ····················  ····················  ····················  ····················
C    681 hCoV-19/England/BRIS-126C96/2020|EPI_ISL_459036|20   ····················  ····················  ····················  ····················  ····················  ····················
C    682 hCoV-19/England/BRIS-126856/2020|EPI_ISL_459038|20   ····················  ····················  ····················  ····················  ····················  ····················
C    683 hCoV-19/England/BRIS-126D57/2020|EPI_ISL_459040|20   ····················  ····················  ····················  ····················  ····················  ····················
C    684 hCoV-19/England/BRIS-1268FC/2020|EPI_ISL_459041|20   ····················  ····················  ····················  ····················  ····················  ····················
C    685 hCoV-19/England/BRIS-126BB7/2020|EPI_ISL_459042|20   ····················  ····················  ····················  ····················  ····················  ····················
C    686 hCoV-19/England/BRIS-126346/2020|EPI_ISL_459046|20   ····················  ····················  ····················  ····················  ····················  ····················
C    687 hCoV-19/England/BRIS-1261B5/2020|EPI_ISL_459047|20   ····················  ····················  ····················  ····················  ····················  ····················
C    688 hCoV-19/England/BRIS-12665C/2020|EPI_ISL_459048|20   ····················  ····················  ····················  ····················  ····················  ····················
C    689 hCoV-19/England/BRIS-126BC6/2020|EPI_ISL_459050|20   ····················  ····················  ····················  ····················  ····················  ····················
C    690 hCoV-19/England/BRIS-126908/2020|EPI_ISL_459051|20   ····················  ·A··················  ····················  ····················  ····················  ····················
C    691 hCoV-19/England/BRIS-1269F9/2020|EPI_ISL_459052|20   ····················  ····················  ····················  ····················  ····················  ····················
C    692 hCoV-19/England/BRIS-126276/2020|EPI_ISL_459053|20   ····················  ····················  ····················  ····················  ····················  ····················
C    693 hCoV-19/England/BRIS-126E45/2020|EPI_ISL_459055|20   ····················  ····················  ····················  ····················  ····················  ····················
C    694 hCoV-19/England/BRIS-1264AD/2020|EPI_ISL_459057|20   ····················  ····················  ····················  ····················  ····················  ····················
C    695 hCoV-19/England/BRIS-125FAD/2020|EPI_ISL_459058|20   ····················  ····················  ····················  ····················  ····················  ····················
C    696 hCoV-19/England/BRIS-126D93/2020|EPI_ISL_459059|20   ····················  ····················  ····················  ····················  ····················  ····················
C    697 hCoV-19/England/BRIS-126100/2020|EPI_ISL_459060|20   ····················  ····················  ····················  ····················  ····················  ····················
C    698 hCoV-19/England/BRIS-1260A9/2020|EPI_ISL_459061|20   ····················  ····················  ····················  ····················  ····················  ····················
C    699 hCoV-19/England/BRIS-1260E5/2020|EPI_ISL_459064|20   ····················  ····················  ····················  ····················  ····················  ····················
C    700 hCoV-19/England/BRIS-126A6F/2020|EPI_ISL_459065|20   ····················  ····················  ····················  ····················  ····················  ····················
C    701 hCoV-19/England/BRIS-1260D6/2020|EPI_ISL_459066|20   ····················  ····················  ····················  ····················  ····················  ····················
C    702 hCoV-19/England/BRIS-12622B/2020|EPI_ISL_459068|20   ····················  ····················  ····················  ····················  ····················  ····················
C    703 hCoV-19/England/BRIS-1265AA/2020|EPI_ISL_459069|20   ····················  ····················  ····················  ····················  ····················  ····················
C    704 hCoV-19/USA/MI-MDHHS-SC20473/2020|EPI_ISL_460012|2   ····················  ····················  ····················  ····················  ····················  ····················
C    705 hCoV-19/USA/MI-MDHHS-SC20476/2020|EPI_ISL_460015|2   ····················  ····················  ····················  ····················  ····················  ····················
C    706 hCoV-19/USA/MI-MDHHS-SC20477/2020|EPI_ISL_460016|2   ····················  ····················  ····················  ····················  ····················  ····················
C    707 hCoV-19/USA/MI-MDHHS-SC20478/2020|EPI_ISL_460017|2   ····················  ····················  ····················  ····················  ····················  ····················
C    708 hCoV-19/USA/MI-MDHHS-SC20479/2020|EPI_ISL_460018|2   ····················  ····················  ····················  ····················  ····················  ····················
```

| | | | | | | | | | |
|---|---|---|---|---|---|---|---|---|---|
| C | 709 hCoV-19/USA/MI-MDHHS-SC20481/2020\|EPI_ISL_460020\|2 | ···················· | ···················· | ···················· | ···················· | ···················· | ···················· | | |
| C | 710 hCoV-19/USA/MI-MDHHS-SC20482/2020\|EPI_ISL_460021\|2 | ···················· | ···················· | ···················· | ···················· | ···················· | ···················· | | |
| C | 711 hCoV-19/USA/MI-MDHHS-SC20484/2020\|EPI_ISL_460023\|2 | ···················· | ···················· | ···················· | ···················· | ···················· | ···················· | | |
| C | 712 hCoV-19/USA/MI-MDHHS-SC20486/2020\|EPI_ISL_460025\|2 | ···················· | ···················· | ···················· | ···················· | ···················· | ···················· | | |
| C | 713 hCoV-19/USA/MI-MDHHS-SC20487/2020\|EPI_ISL_460026\|2 | ···················· | ···················· | ···················· | ···················· | ···················· | ···················· | | |
| C | 714 hCoV-19/USA/MI-MDHHS-SC20489/2020\|EPI_ISL_460028\|2 | ···················· | ···················· | ···················· | ···················· | ···················· | ···················· | | |
| C | 715 hCoV-19/USA/MI-MDHHS-SC20493/2020\|EPI_ISL_460031\|2 | ···················· | ···················· | ···················· | ···················· | ···················· | ···················· | | |
| C | 716 hCoV-19/USA/MI-MDHHS-SC20495/2020\|EPI_ISL_460033\|2 | ···················· | ···················· | ···················· | ···················· | ···················· | ···················· | | |
| C | 717 hCoV-19/USA/MI-MDHHS-SC20496/2020\|EPI_ISL_460034\|2 | ···················· | ···················· | ···················· | ···················· | ···················· | ···················· | | |
| C | 718 hCoV-19/USA/MI-MDHHS-SC20502/2020\|EPI_ISL_460040\|2 | ···················· | ···················· | ···················· | ···················· | ···················· | ···················· | | |
| C | 719 hCoV-19/USA/MI-MDHHS-SC20504/2020\|EPI_ISL_460042\|2 | ···················· | ···················· | ···················· | ···················· | ···················· | ···················· | | |
| C | 720 hCoV-19/USA/MN-MDH-103/2020\|EPI_ISL_460045\|2020-03 | ···················· | ···················· | ···················· | ···················· | ···················· | ···················· | | |
| C | 721 hCoV-19/USA/MN-MDH-104/2020\|EPI_ISL_460046\|2020-03 | ···················· | ···················· | ···················· | ···················· | ···················· | ···················· | | |
| C | 722 hCoV-19/USA/MN-MDH-106/2020\|EPI_ISL_460047\|2020-03 | ···················· | ···················· | ···················· | ···················· | ···················· | ···················· | | |
| C | 723 hCoV-19/USA/MN-MDH-109/2020\|EPI_ISL_460048\|2020-03 | ···················· | ···················· | ···················· | ···················· | ···················· | ···················· | | |
| C | 724 hCoV-19/USA/MN-MDH-123/2020\|EPI_ISL_460049\|2020-03 | ···················· | ···················· | ···················· | ···················· | ···················· | ···················· | | |
| C | 725 hCoV-19/USA/MN-MDH-125/2020\|EPI_ISL_460050\|2020-03 | ···················· | ···················· | ···················· | ···················· | ···················· | ···················· | | |
| C | 726 hCoV-19/USA/MN-MDH-126/2020\|EPI_ISL_460051\|2020-03 | ···················· | ···················· | ···················· | ···················· | ···················· | ···················· | | |
| C | 727 hCoV-19/USA/MN-MDH-139/2020\|EPI_ISL_460052\|2020-03 | ···················· | ···················· | ···················· | ···················· | ···················· | ···················· | | |
| C | 728 hCoV-19/USA/MN-MDH-148/2020\|EPI_ISL_460053\|2020-03 | ···················· | ···················· | ···················· | ···················· | ···················· | ···················· | | |
| C | 729 hCoV-19/USA/MN-MDH-150/2020\|EPI_ISL_460054\|2020-03 | ···················· | ···················· | ···················· | ···················· | ···················· | ···················· | | |
| C | 730 hCoV-19/USA/MA-MGH-00461/2020\|EPI_ISL_460099\|2020- | ···················· | ···················· | ···················· | ···················· | ···················· | ···················· | | |
| C | 731 hCoV-19/Netherlands/ZuidHolland_170/2020\|EPI_ISL_4 | ···················· | ···················· | ···················· | ···················· | ···················· | ···················· | | |
| C | 732 hCoV-19/Netherlands/ZuidHolland_172/2020\|EPI_ISL_4 | ···················· | ···················· | ···················· | ···················· | ···················· | ···················· | | |
| C | 733 hCoV-19/Netherlands/ZuidHolland_173/2020\|EPI_ISL_4 | ···················· | ···················· | ···················· | ···················· | ···················· | ···················· | | |
| C | 734 hCoV-19/Netherlands/ZuidHolland_174/2020\|EPI_ISL_4 | ···················· | ···················· | ···················· | ···················· | ···················· | ···················· | | |
| C | 735 hCoV-19/Netherlands/ZuidHolland_175/2020\|EPI_ISL_4 | ···················· | ···················· | ···················· | ···················· | ···················· | ···················· | | |
| C | 736 hCoV-19/Netherlands/ZuidHolland_176/2020\|EPI_ISL_4 | ···················· | ···················· | ···················· | ···················· | ···················· | ···················· | | |
| C | 737 hCoV-19/Netherlands/ZuidHolland_177/2020\|EPI_ISL_4 | ···················· | ···················· | ···················· | ···················· | ···················· | ···················· | | |
| C | 738 hCoV-19/Netherlands/ZuidHolland_178/2020\|EPI_ISL_4 | ···················· | ···················· | ···················· | ···················· | ···················· | ···················· | | |
| C | 739 hCoV-19/Netherlands/ZuidHolland_179/2020\|EPI_ISL_4 | ···················· | ···················· | ···················· | ···················· | ···················· | ···················· | | |
| C | 740 hCoV-19/Netherlands/ZuidHolland_180/2020\|EPI_ISL_4 | ···················· | ···················· | ···················· | ···················· | ···················· | ···················· | | |
| C | 741 hCoV-19/Netherlands/ZuidHolland_181/2020\|EPI_ISL_4 | ···················· | ···················· | ···················· | ···················· | ···················· | ···················· | | |
| C | 742 hCoV-19/Netherlands/ZuidHolland_183/2020\|EPI_ISL_4 | ···················· | ···················· | ···················· | ···················· | ···················· | ···················· | | |
| C | 743 hCoV-19/Netherlands/ZuidHolland_184/2020\|EPI_ISL_4 | ···················· | ···················· | ···················· | ···················· | ···················· | ···················· | | |
| C | 744 hCoV-19/Netherlands/ZuidHolland_185/2020\|EPI_ISL_4 | ···················· | ···················· | ···················· | ···················· | ···················· | ···················· | | |
| C | 745 hCoV-19/Netherlands/ZuidHolland_186/2020\|EPI_ISL_4 | ···················· | ···················· | ···················· | ···················· | ···················· | ···················· | | |
| C | 746 hCoV-19/Netherlands/ZuidHolland_187/2020\|EPI_ISL_4 | ···················· | ···········A········ | ···················· | ···················· | ···················· | ···················· | | |
| C | 747 hCoV-19/Netherlands/ZuidHolland_188/2020\|EPI_ISL_4 | ···················· | ···················· | ···················· | ···················· | ···················· | ···················· | | |
| C | 748 hCoV-19/Netherlands/ZuidHolland_189/2020\|EPI_ISL_4 | ···················· | ···················· | ···················· | ···················· | ···················· | ···················· | | |
| C | 749 hCoV-19/Netherlands/ZuidHolland_191/2020\|EPI_ISL_4 | ···················· | ···················· | ···················· | ···················· | ···················· | ···················· | | |
| C | 750 hCoV-19/Netherlands/ZuidHolland_192/2020\|EPI_ISL_4 | ···················· | ···················· | ···················· | ···················· | ···················· | ···················· | | |
| C | 751 hCoV-19/Netherlands/ZuidHolland_193/2020\|EPI_ISL_4 | ···················· | ···················· | ···················· | ···················· | ···················· | ···················· | | |
| C | 752 hCoV-19/Netherlands/ZuidHolland_194/2020\|EPI_ISL_4 | ···················· | ···················· | ···················· | ···················· | ···················· | ···················· | | |
| C | 753 hCoV-19/Netherlands/ZuidHolland_195/2020\|EPI_ISL_4 | ···················· | ···················· | ···················· | ···················· | ···················· | ···················· | | |
| C | 754 hCoV-19/Netherlands/ZuidHolland_196/2020\|EPI_ISL_4 | ···················· | ···················· | ···················· | ···················· | ···················· | ···················· | | |
| C | 755 hCoV-19/Netherlands/ZuidHolland_198/2020\|EPI_ISL_4 | ···················· | ···········A········ | ···················· | ···················· | ···················· | ···················· | | |
| C | 756 hCoV-19/Netherlands/ZuidHolland_201/2020\|EPI_ISL_4 | ···················· | ···················· | ···················· | ···················· | ···················· | ···················· | | |
| C | 757 hCoV-19/Netherlands/ZuidHolland_202/2020\|EPI_ISL_4 | ···················· | ···················· | ···················· | ···················· | ···················· | ···················· | | |
| C | 758 hCoV-19/Netherlands/ZuidHolland_203/2020\|EPI_ISL_4 | ···················· | ···················· | ···················· | ···················· | ···················· | ···················· | | |
| C | 759 hCoV-19/Netherlands/ZuidHolland_204/2020\|EPI_ISL_4 | ···················· | ···················· | ···················· | ···················· | ···················· | ···················· | | |
| C | 760 hCoV-19/Netherlands/ZuidHolland_205/2020\|EPI_ISL_4 | ···················· | ···················· | ···················· | ···················· | ···················· | ···················· | | |
| C | 761 hCoV-19/Netherlands/ZuidHolland_206/2020\|EPI_ISL_4 | ···················· | ···················· | ···················· | ···················· | ···················· | ···················· | | |
| C | 762 hCoV-19/Netherlands/ZuidHolland_207/2020\|EPI_ISL_4 | ···················· | ···················· | ···················· | ···················· | ···················· | ···················· | | |
| C | 763 hCoV-19/Netherlands/ZuidHolland_208/2020\|EPI_ISL_4 | ···················· | ···················· | ···················· | ···················· | ···················· | ···················· | | |
| C | 764 hCoV-19/Netherlands/ZuidHolland_209/2020\|EPI_ISL_4 | ···················· | ···················· | ···················· | ···················· | ···················· | ···················· | | |
| C | 765 hCoV-19/Netherlands/ZuidHolland_210/2020\|EPI_ISL_4 | ···················· | ···················· | ···················· | ···················· | ···················· | ···················· | | |
| C | 766 hCoV-19/Netherlands/ZuidHolland_211/2020\|EPI_ISL_4 | ···················· | ···················· | ···················· | ···················· | ···················· | ···················· | | |
| C | 767 hCoV-19/Netherlands/ZuidHolland_212/2020\|EPI_ISL_4 | ···················· | ···················· | ···················· | ···················· | ···················· | ···················· | | |
| C | 768 hCoV-19/Netherlands/ZuidHolland_213/2020\|EPI_ISL_4 | ···················· | ···················· | ···················· | ···················· | ···················· | ···················· | | |
| C | 769 hCoV-19/England/NORW-E8CDD/2020\|EPI_ISL_448300\|202 | ···················· | ···················· | ···················· | ···················· | ···················· | ···················· | | |
| C | 770 hCoV-19/England/NORW-E8CEC/2020\|EPI_ISL_448301\|202 | ···················· | ···················· | ···················· | ···················· | ···················· | ···················· | | |
| C | 771 hCoV-19/England/NORW-E8D16/2020\|EPI_ISL_448304\|202 | ···················· | ···················· | ···················· | ···················· | ···················· | ···················· | | |
| C | 772 hCoV-19/England/NORW-E8D34/2020\|EPI_ISL_448305\|202 | ···················· | ···················· | ···················· | ···················· | ···················· | ···················· | | |
| C | 773 hCoV-19/England/NORW-E8D52/2020\|EPI_ISL_448306\|202 | ···················· | ···················· | ···················· | ···················· | ···················· | ···················· | | |
| C | 774 hCoV-19/England/NORW-E8D61/2020\|EPI_ISL_448307\|202 | ···················· | ···················· | ···················· | ···················· | ···················· | ···················· | | |
| C | 775 hCoV-19/England/NORW-E8D70/2020\|EPI_ISL_448308\|202 | ···················· | ···················· | ···················· | ···················· | ···················· | ···················· | | |
| C | 777 hCoV-19/England/NORW-E9401/2020\|EPI_ISL_448311\|202 | ··················T· | ···················· | ···················· | ···················· | ···················· | ···················· | 1 | |
| C | 778 hCoV-19/England/NORW-E942F/2020\|EPI_ISL_448312\|202 | ···················· | ···················· | ···················· | ···················· | ···················· | ···················· | | |
| C | 779 hCoV-19/England/NORW-E944D/2020\|EPI_ISL_448313\|202 | ···················· | ···················· | ···················· | ···················· | ···················· | ···················· | | |
| C | 780 hCoV-19/England/NORW-E946B/2020\|EPI_ISL_448314\|202 | ···················· | ···················· | ···················· | ···················· | ···················· | ···················· | | |
| C | 781 hCoV-19/England/NORW-E9498/2020\|EPI_ISL_448315\|202 | ···················· | ···················· | ···················· | ···················· | NNN················· | ···················· | | 1 |
| C | 782 hCoV-19/England/NORW-E94A7/2020\|EPI_ISL_448316\|202 | ···················· | ···················· | ···················· | ···················· | ···················· | ···················· | | |
| C | 783 hCoV-19/England/NORW-E94F2/2020\|EPI_ISL_448317\|202 | ···················· | ···················· | ···················· | ···················· | ·NN················· | ···················· | | |
| C | 784 hCoV-19/England/NORW-E954A/2020\|EPI_ISL_448320\|202 | ···················· | ···················· | ···················· | ···················· | ···················· | ···················· | | |
| C | 785 hCoV-19/England/NORW-E95A4/2020\|EPI_ISL_448321\|202 | ··················T· | ···················· | ···················· | ···················· | ···················· | ···················· | 1 | |
| C | 786 hCoV-19/England/NORW-E960B/2020\|EPI_ISL_448322\|202 | ···················· | ···················· | ···················· | ···················· | ···················· | ···················· | | |
| C | 788 hCoV-19/England/NORW-E9665/2020\|EPI_ISL_448324\|202 | ···················· | ···················A | ···················· | ···················· | ···················· | ···················· | 1 | |

| | | | | | | | | |
|---|---|---|---|---|---|---|---|---|
| C | 790 hCoV-19/England/NORW-E96B0/2020\|EPI_ISL_448326\|202 | .................... | .................... | .................... | .................... | .................... | .................... | |
| C | 791 hCoV-19/England/NORW-E96CF/2020\|EPI_ISL_448327\|202 | .................... | ...................A | .................... | .................... | .................... | .................... | 1 |
| C | 792 hCoV-19/England/NORW-E9744/2020\|EPI_ISL_448328\|202 | .................... | .................... | .................... | .................... | .................... | .................... | |
| C | 793 hCoV-19/England/NORW-E9753/2020\|EPI_ISL_448329\|202 | .................... | .................... | .................... | .................... | .................... | .................... | |
| C | 795 hCoV-19/England/NORW-E97AE/2020\|EPI_ISL_448331\|202 | .................... | .................... | .................... | .................... | .................... | .................... | |
| C | 796 hCoV-19/England/NORW-E97BD/2020\|EPI_ISL_448332\|202 | .................... | .................... | .................... | .................... | .................... | .................... | |
| C | 797 hCoV-19/England/NORW-E9805/2020\|EPI_ISL_448335\|202 | .................... | .................... | .................... | .................... | .................... | .................... | |
| C | 798 hCoV-19/England/NORW-E9814/2020\|EPI_ISL_448336\|202 | .................... | .................... | .................... | .................... | .................... | .................... | |
| C | 799 hCoV-19/England/NORW-E9841/2020\|EPI_ISL_448338\|202 | .................... | .................... | .................... | .................... | .................... | .................... | |
| C | 800 hCoV-19/England/NORW-E987E/2020\|EPI_ISL_448339\|202 | .................... | ...................A | .................... | .................... | .................... | .................... | 1 |
| C | 801 hCoV-19/England/NORW-E9CEB/2020\|EPI_ISL_448360\|202 | ..................T. | .................... | .................... | .................... | .................... | .................... | 1 |
| C | 802 hCoV-19/England/NORW-E9D06/2020\|EPI_ISL_448361\|202 | .................... | .................... | .................... | .................... | .................... | .................... | |
| C | 803 hCoV-19/England/NORW-E9D42/2020\|EPI_ISL_448362\|202 | .................... | .................... | .................... | .................... | .................... | .................... | |
| C | 804 hCoV-19/England/NORW-E9D60/2020\|EPI_ISL_448363\|202 | .................... | .................... | .................... | .................... | .................... | .................... | |
| C | 805 hCoV-19/England/NORW-E9D7F/2020\|EPI_ISL_448364\|202 | .................... | .................... | .................... | .................... | .................... | .................... | |
| C | 806 hCoV-19/England/NORW-E9D8E/2020\|EPI_ISL_448365\|202 | .................... | .................... | .................... | .................... | .................... | .................... | |
| C | 807 hCoV-19/England/NORW-E9D9D/2020\|EPI_ISL_448366\|202 | .................... | .................... | .................... | .................... | .................... | .................... | |
| C | 808 hCoV-19/England/NORW-E9DE8/2020\|EPI_ISL_448369\|202 | .................... | .................... | .................... | .................... | .................... | .................... | |
| C | 809 hCoV-19/England/NORW-E9DF7/2020\|EPI_ISL_448370\|202 | .................... | .................... | .................... | .................... | .................... | .................... | |
| C | 810 hCoV-19/England/NORW-E9E12/2020\|EPI_ISL_448371\|202 | .................... | .................... | .................... | .................... | .................... | .................... | |
| C | 811 hCoV-19/England/NORW-E9E30/2020\|EPI_ISL_448372\|202 | .................... | .................... | .................... | .................... | .................... | .................... | |
| C | 812 hCoV-19/England/NORW-E9E6D/2020\|EPI_ISL_448374\|202 | .................... | .................... | .................... | .................... | .................... | .................... | |
| C | 813 hCoV-19/England/NORW-E9E7C/2020\|EPI_ISL_448375\|202 | .................... | .................... | .................... | .................... | .................... | .................... | |
| C | 814 hCoV-19/England/NORW-E9E9A/2020\|EPI_ISL_448376\|202 | .................... | .................... | .................... | .................... | .................... | .................... | |
| C | 815 hCoV-19/England/NORW-E9EA9/2020\|EPI_ISL_448377\|202 | .................... | .................... | .................... | .................... | .................... | .................... | |
| C | 817 hCoV-19/England/LOND-D3B14/2020\|EPI_ISL_449627\|202 | .................... | .................... | .................... | .................... | .................... | .................... | |
| C | 818 hCoV-19/England/LOND-D3B32/2020\|EPI_ISL_449629\|202 | .................... | .................... | .................... | .................... | .................... | .................... | |
| C | 819 hCoV-19/England/LOND-D3B41/2020\|EPI_ISL_449630\|202 | .................... | .................... | .................... | .................... | .................... | .................... | |
| C | 820 hCoV-19/England/LOND-D3B50/2020\|EPI_ISL_449631\|202 | .................... | .................... | .................... | .................... | .................... | .................... | |
| C | 821 hCoV-19/England/LOND-D3B6F/2020\|EPI_ISL_449632\|202 | .................... | .................... | .................... | .................... | .................... | .................... | |
| C | 822 hCoV-19/England/LOND-D3B8D/2020\|EPI_ISL_449633\|202 | .................... | .................... | .................... | .................... | .................... | .................... | |
| C | 823 hCoV-19/England/LOND-D3BBA/2020\|EPI_ISL_449636\|202 | .................... | .................... | .................... | .................... | .................... | .................... | |
| C | 824 hCoV-19/England/LOND-D3BC9/2020\|EPI_ISL_449637\|202 | .................... | .................... | .................... | .................... | .................... | .................... | |
| C | 825 hCoV-19/England/LOND-D3BE7/2020\|EPI_ISL_449639\|202 | .................... | .................... | .................... | .................... | .................... | .................... | |
| C | 826 hCoV-19/England/LOND-D3BF6/2020\|EPI_ISL_449640\|202 | ...T................ | .................... | .................... | .................... | .................... | .................... | |
| C | 827 hCoV-19/England/LOND-D3C3F/2020\|EPI_ISL_449644\|202 | .................... | .................... | .................... | .................... | .................... | .................... | |
| C | 828 hCoV-19/England/LOND-D3D3C/2020\|EPI_ISL_449645\|202 | .................... | .................... | .................... | .................... | .................... | .................... | |
| C | 829 hCoV-19/England/LOND-D3D4B/2020\|EPI_ISL_449646\|202 | .................... | .................... | .................... | .................... | .................... | .................... | |
| C | 830 hCoV-19/England/LOND-D3D5A/2020\|EPI_ISL_449647\|202 | .................... | .................... | .................... | .................... | .................... | .................... | |
| C | 831 hCoV-19/England/LOND-D3D69/2020\|EPI_ISL_449648\|202 | .................... | .................... | .................... | .................... | .................... | .................... | |
| C | 832 hCoV-19/England/LOND-D3D78/2020\|EPI_ISL_449649\|202 | .................... | .................... | .................... | .................... | .................... | .................... | |
| C | 833 hCoV-19/England/LOND-D3D87/2020\|EPI_ISL_449650\|202 | .................... | .................... | .................... | .................... | .................... | .................... | |
| C | 834 hCoV-19/England/LOND-D3D96/2020\|EPI_ISL_449651\|202 | .................... | .................... | .................... | .................... | .................... | .................... | |
| C | 835 hCoV-19/England/LOND-D3DA5/2020\|EPI_ISL_449652\|202 | .................... | .................... | .................... | .................... | .................... | .................... | |
| C | 836 hCoV-19/England/LOND-D3DB4/2020\|EPI_ISL_449653\|202 | .................... | .................... | .................... | .................... | .................... | .................... | |
| C | 837 hCoV-19/England/LOND-D3DC3/2020\|EPI_ISL_449654\|202 | .................... | .................... | .................... | .................... | .................... | .................... | |
| C | 838 hCoV-19/England/LOND-D3DD2/2020\|EPI_ISL_449655\|202 | .................... | .................... | .................... | .................... | .................... | .................... | |
| C | 839 hCoV-19/England/LOND-D4B9B/2020\|EPI_ISL_449656\|202 | .................... | .................... | .................... | .................... | .................... | .................... | |
| C | 840 hCoV-19/England/LOND-D4BAA/2020\|EPI_ISL_449657\|202 | .................... | .................... | .................... | .................... | .................... | .................... | |
| C | 841 hCoV-19/England/LOND-D4BB9/2020\|EPI_ISL_449658\|202 | .................... | .................... | .................... | .................... | .................... | .................... | |
| C | 842 hCoV-19/England/LOND-D4BC8/2020\|EPI_ISL_449659\|202 | .................... | .................... | .................... | .................... | .................... | .................... | |
| C | 843 hCoV-19/England/LOND-D4BD7/2020\|EPI_ISL_449660\|202 | .................... | .................... | .................... | .................... | .................... | .................... | |
| C | 844 hCoV-19/England/LOND-D4BE6/2020\|EPI_ISL_449661\|202 | .................... | .................... | .................... | .................... | .................... | .................... | |
| C | 845 hCoV-19/England/LOND-D4BF5/2020\|EPI_ISL_449662\|202 | .................... | .................... | .................... | .................... | .................... | .................... | |
| C | 846 hCoV-19/England/LOND-D4C01/2020\|EPI_ISL_449663\|202 | .................... | .................... | .................... | .................... | .................... | .................... | |
| C | 847 hCoV-19/England/LOND-D4C10/2020\|EPI_ISL_449664\|202 | .................... | .................... | .................... | .................... | .................... | .................... | |
| C | 848 hCoV-19/England/LOND-D4C2F/2020\|EPI_ISL_449665\|202 | .................... | .................... | .................... | .................... | .................... | .................... | |
| C | 849 hCoV-19/England/LOND-D4C3E/2020\|EPI_ISL_449666\|202 | .................... | .................... | .................... | .................... | .................... | .................... | |
| C | 850 hCoV-19/England/LOND-D4C4D/2020\|EPI_ISL_449667\|202 | .................... | .................... | .................... | .................... | .................... | .................... | |
| C | 851 hCoV-19/England/LOND-D4C5C/2020\|EPI_ISL_449668\|202 | .................... | .................... | .................... | .................... | .................... | .................... | |
| C | 852 hCoV-19/England/LOND-D4C6B/2020\|EPI_ISL_449669\|202 | .................... | .................... | .................... | .................... | .................... | .................... | |
| C | 853 hCoV-19/England/LOND-D521D/2020\|EPI_ISL_449690\|202 | .................... | .................... | .................... | .................... | .................... | .................... | |
| C | 854 hCoV-19/England/LOND-D522C/2020\|EPI_ISL_449691\|202 | .................... | .................... | .................... | .................... | .................... | .................... | |
| C | 855 hCoV-19/England/LOND-D523B/2020\|EPI_ISL_449692\|202 | .................... | .................... | .................... | .................... | .................... | .................... | |
| C | 856 hCoV-19/England/LOND-D5268/2020\|EPI_ISL_449694\|202 | .................... | .................... | .................... | .................... | .................... | .................... | |
| C | 857 hCoV-19/England/LOND-D5286/2020\|EPI_ISL_449696\|202 | .................... | .................... | .................... | .................... | .................... | .................... | |
| C | 858 hCoV-19/England/LOND-D559C/2020\|EPI_ISL_449698\|202 | .................... | .................... | .................... | .................... | .................... | .................... | |
| C | 859 hCoV-19/USA/MI-MDHHS-SC20331/2020\|EPI_ISL_450610\|2 | .................... | .................... | .................... | .................... | .................... | .................... | |
| C | 860 hCoV-19/USA/MI-MDHHS-SC20333/2020\|EPI_ISL_450612\|2 | .................... | .................... | .................... | .................... | .................... | .................... | |
| C | 861 hCoV-19/USA/MI-MDHHS-SC20335/2020\|EPI_ISL_450614\|2 | .................... | .................... | .................... | .................... | .................... | .................... | |
| C | 862 hCoV-19/USA/MI-MDHHS-SC20337/2020\|EPI_ISL_450616\|2 | .................... | .................... | .................... | .................... | .................... | .................... | |
| C | 863 hCoV-19/USA/MI-MDHHS-SC20339/2020\|EPI_ISL_450618\|2 | .................... | .................... | .................... | .................... | .................... | .................... | |
| C | 864 hCoV-19/USA/MI-MDHHS-SC20341/2020\|EPI_ISL_450620\|2 | .................... | .................... | .................... | .................... | .................... | .................... | |
| C | 865 hCoV-19/USA/MI-MDHHS-SC20344/2020\|EPI_ISL_450623\|2 | .................... | .................... | .................... | .................... | .................... | .................... | |
| C | 866 hCoV-19/USA/MI-MDHHS-SC20352/2020\|EPI_ISL_450631\|2 | .................... | .................... | .................... | .................... | .................... | .................... | |
| C | 867 hCoV-19/USA/MI-MDHHS-SC20356/2020\|EPI_ISL_450635\|2 | .................... | .................... | .................... | .................... | .................... | .................... | |
| C | 868 hCoV-19/USA/MI-MDHHS-SC20357/2020\|EPI_ISL_450636\|2 | .................... | .................... | .................... | .................... | .................... | .................... | |
| C | 869 hCoV-19/Canada/QC-AJ7/2020\|EPI_ISL_450639\|2020-04- | .................... | .................... | .................... | .................... | .................... | .................... | |

| | | | | | | | | |
|---|---|---|---|---|---|---|---|---|
| C | 870 hCoV-19/Germany/BY-MVP-0123/2020\|EPI_ISL_451940\|20 | ···················· | ···················· | ···················· | ···················· | ···················· | ···················· | |
| C | 871 hCoV-19/Germany/BY-MVP-0124/2020\|EPI_ISL_451941\|20 | ···················· | ···················· | ···················· | ···················· | T··················· | ···················· | |
| C | 872 hCoV-19/Germany/BY-MVP-0153/2020\|EPI_ISL_451944\|20 | ···················· | ···················· | ···················· | ···················· | ···················· | ···················· | |
| C | 873 hCoV-19/Germany/BY-MVP-0155/2020\|EPI_ISL_451945\|20 | ···················· | ···················· | ···················· | ···················· | ···················· | ···················· | |
| C | 874 hCoV-19/Belarus/ChVir2070/2020\|EPI_ISL_451948\|2020 | ···················· | ······C············· | ···················· | ···················· | ···················· | ···················· | |
| C | 875 hCoV-19/Pakistan/KHI1/2020\|EPI_ISL_451958\|2020-03- | ···················· | ···················· | ···················· | ···················· | ···················· | ···················· | |
| C | 876 hCoV-19/USA/AK-UAF-087/2020\|EPI_ISL_451959\|2020-03 | ···················· | ···················· | ···················· | ···················· | ···················· | ···················· | |
| C | 877 hCoV-19/Italy/APU-UniMI-CL1/2020\|EPI_ISL_451961\|20 | ···················· | ···················· | ···················· | ···················· | ···················· | ···················· | |
| C | 878 hCoV-19/Russia/SCPM-O-01/2020\|EPI_ISL_451963\|2020- | ···················· | ···················· | ···················· | ···················· | ···················· | ···················· | |
| C | 879 hCoV-19/Russia/SCPM-O-04/2020\|EPI_ISL_451966\|2020- | ···················· | ···················· | ···················· | ···················· | ···················· | ···················· | |
| C | 880 hCoV-19/Russia/SCPM-O-07/2020\|EPI_ISL_451969\|2020- | ···················· | ···················· | ···················· | ···················· | ···················· | ···················· | |
| C | 881 hCoV-19/Russia/SCPM-O-08/2020\|EPI_ISL_451970\|2020- | ···················· | ···················· | ···················· | ···················· | ···················· | ···················· | |
| C | 882 hCoV-19/Poland/PL_P10/2020\|EPI_ISL_451971\|2020-04- | ···················· | ···················· | ···················· | ···················· | ···················· | ···················· | |
| C | 883 hCoV-19/Poland/PL_P11/2020\|EPI_ISL_451972\|2020-04- | ···················· | ···················· | ···················· | ···················· | ···················· | ···················· | |
| C | 884 hCoV-19/Poland/PL_P12/2020\|EPI_ISL_451973\|2020-03- | ···················· | ···················· | ···················· | ···················· | ···················· | ···················· | |
| C | 885 hCoV-19/Poland/PL_P13/2020\|EPI_ISL_451974\|2020-04- | ···················· | ···················· | ···················· | ···················· | ···················· | ···················· | |
| C | 886 hCoV-19/Poland/PL_P14/2020\|EPI_ISL_451975\|2020-03- | ···················· | ···················· | ···················· | ···················· | ···················· | ···················· | |
| C | 887 hCoV-19/Poland/PL_P15/2020\|EPI_ISL_451976\|2020-04- | ···················· | ···················· | ···················· | ···················· | ···················· | ···················· | |
| C | 888 hCoV-19/Poland/PL_P16/2020\|EPI_ISL_451977\|2020-04- | ···················· | ···················· | ···················· | ···················· | ···················· | ···················· | |
| C | 889 hCoV-19/Poland/PL_P17/2020\|EPI_ISL_451978\|2020-04- | ···················· | ···················· | ···················· | ···················· | ···················· | ···················· | |
| C | 890 hCoV-19/Poland/PL_P18/2020\|EPI_ISL_451979\|2020-03- | ···················· | ···················· | ···················· | ···················· | ···················· | ···················· | |
| C | 891 hCoV-19/Poland/PL_P19/2020\|EPI_ISL_451980\|2020-04- | ···················· | ···················· | ···················· | ···················· | ···················· | ···················· | |
| C | 892 hCoV-19/Scotland/CVR3371/2020\|EPI_ISL_456801\|2020- | ···················· | ···················· | ···················· | ···················· | ···················· | ···················· | |
| C | 893 hCoV-19/Scotland/CVR3373/2020\|EPI_ISL_456802\|2020- | ···················· | ···················· | ···················· | ···················· | ···················· | ···················· | |
| C | 894 hCoV-19/Scotland/CVR3374/2020\|EPI_ISL_456803\|2020- | ···················· | ···················A | ···················· | ···················· | ···················· | ···················· | 1 |
| C | 895 hCoV-19/Scotland/CVR3375/2020\|EPI_ISL_456804\|2020- | ···················· | ···················· | ···················· | ···················· | ···················· | ···················· | |
| C | 896 hCoV-19/Scotland/CVR3376/2020\|EPI_ISL_456805\|2020- | ···················· | ···················· | ···················· | ···················· | ···················· | ···················· | |
| C | 897 hCoV-19/Scotland/CVR3377/2020\|EPI_ISL_456806\|2020- | ···················· | ···················· | ···················· | ···················· | ···················· | ···················· | |
| C | 898 hCoV-19/Scotland/CVR3378/2020\|EPI_ISL_456807\|2020- | ···················· | ···················· | ···················· | ···················· | ···················· | ···················· | |
| C | 899 hCoV-19/Scotland/CVR3379/2020\|EPI_ISL_456808\|2020- | ···················· | ···················· | ···················· | ···················· | ···················· | ···················· | |
| C | 900 hCoV-19/Scotland/CVR3380/2020\|EPI_ISL_456809\|2020- | ···················· | ···················A | ···················· | ···················· | ···················· | ···················· | 1 |
| C | 901 hCoV-19/Scotland/CVR3381/2020\|EPI_ISL_456810\|2020- | ···················· | ···················A | ···················· | ···················· | ···················· | ···················· | 1 |
| C | 903 hCoV-19/Scotland/CVR3385/2020\|EPI_ISL_456813\|2020- | ···················· | ···················· | ···················· | ···················· | ···················· | ···················· | |
| C | 906 hCoV-19/Scotland/CVR3392/2020\|EPI_ISL_456819\|2020- | ···················· | ···················· | ···················· | ···················· | ···················· | ···················· | |
| C | 910 hCoV-19/Scotland/CVR3402/2020\|EPI_ISL_456827\|2020- | ···················· | ···················A | ···················· | ···················· | ···················· | ···················· | 1 |
| C | 911 hCoV-19/Scotland/CVR3405/2020\|EPI_ISL_456829\|2020- | ···················· | ···················· | ···················· | ···················· | ···················· | ···················· | |
| C | 912 hCoV-19/Scotland/CVR3406/2020\|EPI_ISL_456830\|2020- | ···················· | ···················A | ···················· | ···················· | ···················· | ···················· | 1 |
| C | 914 hCoV-19/England/BRIS-126470/2020\|EPI_ISL_459014\|20 | ···················· | ···················· | ···················· | ···················· | ···················· | ···················· | |
| C | 915 hCoV-19/England/BRIS-126795/2020\|EPI_ISL_459015\|20 | ···················· | ···················· | ···················· | ···················· | ···················· | ···················· | |
| C | 916 hCoV-19/England/BRIS-126C5A/2020\|EPI_ISL_459016\|20 | ···················· | ···················· | ···················· | ···················· | ···················· | ···················· | |
| C | 917 hCoV-19/England/BRIS-125F9E/2020\|EPI_ISL_459020\|20 | ···················· | ···················· | ···················· | ···················· | ···················· | ···················· | |
| C | 918 hCoV-19/England/BRIS-125FDA/2020\|EPI_ISL_459021\|20 | ···················· | ···················· | ···················· | ···················· | ···················· | ···················· | |
| C | 919 hCoV-19/England/BRIS-126003/2020\|EPI_ISL_459022\|20 | ···················· | ···················· | ···················· | ···················· | ···················· | ···················· | |
| C | 920 hCoV-19/England/BRIS-12674A/2020\|EPI_ISL_459024\|20 | ···················· | ···················· | ···················· | ···················· | ···················· | ···················· | |
| C | 921 hCoV-19/England/BRIS-1263DD/2020\|EPI_ISL_459025\|20 | ···················· | ···················· | ···················· | ···················· | ···················· | ···················· | |
| C | 922 hCoV-19/England/BRIS-1262B2/2020\|EPI_ISL_459026\|20 | ···················· | ····A··············· | ···················· | ···················· | ···················· | ···················· | |
| C | 923 hCoV-19/England/BRIS-12671D/2020\|EPI_ISL_459027\|20 | ···················· | ···················· | ···················· | ···················· | ···················· | ···················· | |
| C | 924 hCoV-19/England/BRIS-126A8D/2020\|EPI_ISL_459028\|20 | ···················· | ···················· | ···················· | ···················· | ···················· | ···················· | |
| C | 925 hCoV-19/England/BRIS-12621C/2020\|EPI_ISL_459029\|20 | ···················· | ···················· | ···················· | ···················· | ···················· | ···················· | |
| C | 926 hCoV-19/England/BRIS-126689/2020\|EPI_ISL_459030\|20 | ···················· | ···················· | ···················· | ···················· | ···················· | ···················· | |
| C | 927 hCoV-19/England/BRIS-126B20/2020\|EPI_ISL_459033\|20 | ···················· | ···················· | ···················· | ···················· | ···················· | ···················· | |
| C | 928 hCoV-19/England/BRIS-12699F/2020\|EPI_ISL_459034\|20 | ···················· | ···················· | ···················· | ···················· | ···················· | ···················· | |
| C | 929 hCoV-19/England/BRIS-1263CE/2020\|EPI_ISL_459070\|20 | ···················· | ···················· | ···················· | ···················· | ···················· | ···················· | |
| C | 930 hCoV-19/England/BRIS-1264F8/2020\|EPI_ISL_459071\|20 | ···················· | ···················· | ···················· | ···················· | ···················· | ···················· | |
| C | 931 hCoV-19/England/BRIS-126D48/2020\|EPI_ISL_459076\|20 | ···················· | ···················· | ···················· | ···················· | ···················· | ···················· | |
| C | 932 hCoV-19/England/BRIS-12608B/2020\|EPI_ISL_459077\|20 | ···················· | ···················· | ···················· | ···················· | ···················· | ···················· | |
| C | 933 hCoV-19/England/BRIS-1265B9/2020\|EPI_ISL_459078\|20 | ···················· | ···················· | ···················· | ···················· | ···················· | ···················· | |
| C | 934 hCoV-19/England/BRIS-126C1E/2020\|EPI_ISL_459079\|20 | ···················· | ···················· | ···················· | ···················· | ···················· | ···················· | |
| C | 935 hCoV-19/England/BRIS-1265C8/2020\|EPI_ISL_459080\|20 | ···················· | ···················· | ···················· | ···················· | ···················· | ···················· | |
| C | 936 hCoV-19/England/BRIS-12616A/2020\|EPI_ISL_459081\|20 | ···················· | ·············A······ | ···················· | ···················· | ···················· | ···················· | |
| C | 937 hCoV-19/England/BRIS-126267/2020\|EPI_ISL_459082\|20 | ···················· | ···················· | ···················· | ···················· | ···················· | ···················· | |
| C | 938 hCoV-19/England/BRIS-126AD8/2020\|EPI_ISL_459083\|20 | ···················· | ···················· | ···················· | ···················· | ···················· | ···················· | |
| C | 939 hCoV-19/England/BRIS-12609A/2020\|EPI_ISL_459085\|20 | ···················· | ···················· | ···················· | ···················· | ···················· | ···················· | |
| C | 940 hCoV-19/England/BRIS-126C69/2020\|EPI_ISL_459086\|20 | ···················· | ···················· | ···················· | ···················· | ···················· | ···················· | |
| C | 941 hCoV-19/England/BRIS-126A9C/2020\|EPI_ISL_459090\|20 | ···················· | ···················· | ···················· | ···················· | ···················· | ···················· | |
| C | 942 hCoV-19/England/BRIS-126874/2020\|EPI_ISL_459091\|20 | ···················· | ···················· | ···················· | ···················· | ···················· | ···················· | |
| C | 943 hCoV-19/England/BRIS-1260C7/2020\|EPI_ISL_459093\|20 | ···················· | ···················· | ···················· | ···················· | ···················· | ···················· | |
| C | 944 hCoV-19/Scotland/GCVR-172B70/2020\|EPI_ISL_459096\|2 | ···················· | ···················· | ···················· | ···················· | ···················· | ···················· | |
| C | 945 hCoV-19/Scotland/GCVR-173A63/2020\|EPI_ISL_459097\|2 | ···················· | ···········A········ | ···················· | ···················· | ···················· | ···················· | |
| C | 946 hCoV-19/Scotland/GCVR-173C30/2020\|EPI_ISL_459098\|2 | ···················· | ···················· | ···················· | ···················· | ···················· | ···················· | |
| C | 947 hCoV-19/Scotland/GCVR-173B9D/2020\|EPI_ISL_459099\|2 | ···················· | ···················· | ···················· | ···················· | ···················· | ···················· | |
| C | 948 hCoV-19/USA/MI-MDHHS-SC20464/2020\|EPI_ISL_460003\|2 | ···················· | ···················· | ···················· | ···················· | ···················· | ···················· | |
| C | 949 hCoV-19/USA/MI-MDHHS-SC20465/2020\|EPI_ISL_460004\|2 | ···················· | ···················· | ···················· | ···················· | ···················· | ···················· | |
| C | 950 hCoV-19/USA/MI-MDHHS-SC20469/2020\|EPI_ISL_460008\|2 | ···················· | ···················· | ···················· | ···················· | ···················· | ···················· | |
| C | 951 hCoV-19/USA/MI-MDHHS-SC20470/2020\|EPI_ISL_460009\|2 | ···················· | ···················· | ···················· | ···················· | ···················· | ···················· | |
| C | 952 hCoV-19/USA/MN-MDH-152/2020\|EPI_ISL_460055\|2020-03 | ···················· | ···················· | ···················· | ···················· | ···················· | ···················· | |
| C | 953 hCoV-19/USA/MN-MDH-156/2020\|EPI_ISL_460056\|2020-03 | ···················· | ···················· | ···················· | ···················· | ···················· | ···················· | |
| C | 954 hCoV-19/USA/MN-MDH-162/2020\|EPI_ISL_460057\|2020-03 | ···················· | ···················· | ···················· | ···················· | ···················· | ···················· | |

| | | | | | | | | |
|---|---|---|---|---|---|---|---|---|
| C | 955 hCoV-19/USA/MN-MDH-163/2020\|EPI_ISL_460058\|2020-03 | ···················· | ···················· | ···················· | ···················· | ···················· | ···················· | |
| C | 956 hCoV-19/USA/MN-MDH-164/2020\|EPI_ISL_460059\|2020-03 | ···················· | ···················· | ···················· | ···················· | ···················· | ···················· | |
| C | 957 hCoV-19/USA/MN-MDH-70/2020\|EPI_ISL_460060\|2020-03- | ···················· | ···················· | ···················· | ···················· | ···················· | ···················· | |
| C | 958 hCoV-19/USA/MN-MDH-71/2020\|EPI_ISL_460061\|2020-03- | ···················· | ···················· | ···················· | ···················· | ···················· | ···················· | |
| C | 959 hCoV-19/USA/MN-MDH-72/2020\|EPI_ISL_460062\|2020-03- | ···················· | ···················· | ···················· | ···················· | ···················· | ···················· | |
| C | 960 hCoV-19/USA/MN-MDH-74/2020\|EPI_ISL_460063\|2020-03- | ···················· | ···················· | ···················· | ···················· | ···················· | ···················· | |
| C | 961 hCoV-19/USA/MN-MDH-75/2020\|EPI_ISL_460064\|2020-03- | ···················· | ···················· | ···················· | ···················· | ···················· | ···················· | |
| C | 962 hCoV-19/USA/MN-MDH-78/2020\|EPI_ISL_460065\|2020-03- | ···················· | ···················· | ···················· | ···················· | ···················· | ···················· | |
| C | 963 hCoV-19/USA/MN-MDH-79/2020\|EPI_ISL_460066\|2020-03- | ···················· | ···················· | ···················· | ···················· | ···················· | ···················· | |
| C | 964 hCoV-19/USA/MN-MDH-83/2020\|EPI_ISL_460067\|2020-03- | ···················· | ···················· | ···················· | ···················· | ···················· | ···················· | |
| C | 965 hCoV-19/USA/MN-MDH-84/2020\|EPI_ISL_460068\|2020-03- | ···················· | ···················· | ···················· | ···················· | ···················· | ···················· | |
| C | 966 hCoV-19/USA/MN-MDH-85/2020\|EPI_ISL_460069\|2020-03- | ···················· | ···················· | ···················· | ···················· | ···················· | ···················· | |
| C | 967 hCoV-19/USA/MN-MDH-87/2020\|EPI_ISL_460070\|2020-03- | ···················· | ···················· | ···················· | ···················· | ···················· | ···················· | |
| C | 968 hCoV-19/USA/MN-MDH-89/2020\|EPI_ISL_460071\|2020-03- | ···················· | ···················· | ···················· | ···················· | ···················· | ···················· | |
| C | 969 hCoV-19/USA/MN-MDH-91/2020\|EPI_ISL_460072\|2020-03- | ···················· | ···················· | ···················· | ···················· | ···················· | ···················· | |
| C | 970 hCoV-19/USA/MN-MDH-92/2020\|EPI_ISL_460073\|2020-03- | ···················· | ···················· | ···················· | ···················· | ···················· | ···················· | |
| C | 971 hCoV-19/USA/MN-MDH-94/2020\|EPI_ISL_460075\|2020-03- | ···················· | ···················· | ···················· | ···················· | ···················· | ···················· | |
| C | 972 hCoV-19/USA/MN-MDH-95/2020\|EPI_ISL_460076\|2020-03- | ···················· | ···················· | ···················· | ···················· | ···················· | ···················· | |
| C | 973 hCoV-19/USA/MN-MDH-97/2020\|EPI_ISL_460077\|2020-03- | ···················· | ···················· | ···················· | ···················· | ···················· | ···················· | |
| C | 974 hCoV-19/USA/MN-MDH-98/2020\|EPI_ISL_460078\|2020-03- | ···················· | ···················· | ···················· | ···················· | ···················· | ···················· | |
| C | 975 hCoV-19/Italy/LOM-INMI-BG-5825/2020\|EPI_ISL_460079 | ···················· | ···················· | ···················· | ···················· | ···················· | ···················· | |
| C | 976 hCoV-19/Italy/LOM-INMI-BG-11639/2020\|EPI_ISL_46008 | ···················· | ···················· | ···················· | ···················· | ···················· | ···················· | |
| C | 977 hCoV-19/Italy/LOM-INMI-BG-7158/2020\|EPI_ISL_460081 | ···················· | ···················· | ···················· | ···················· | ···················· | ···················· | |
| C | 978 hCoV-19/Italy/LOM-INMI-BG-9209/2020\|EPI_ISL_460082 | ···················· | ···················· | ···················· | ···················· | ···················· | ···················· | |
| C | 979 hCoV-19/Italy/LOM-INMI-BG-9225/2020\|EPI_ISL_460083 | ···················· | ···················· | ···················· | ···················· | ···················· | ···················· | |
| C | 980 hCoV-19/Italy/LOM-INMI-BS-12256/2020\|EPI_ISL_46008 | ···················· | ···················· | ···················· | ···················· | ···················· | ···················· | |
| C | 981 hCoV-19/Italy/LOM-INMI-13221/2020\|EPI_ISL_460085\|2 | ···················· | ···················· | ···················· | ···················· | ···················· | ···················· | |
| C | 982 hCoV-19/Italy/LOM-INMI-7070/2020\|EPI_ISL_460086\|20 | ···················· | ···················· | ···················· | ···················· | ···················· | ···················· | |
| C | 983 hCoV-19/Italy/LOM-INMI-9206/2020\|EPI_ISL_460087\|20 | ···················· | ···················· | ···················· | ···················· | ···················· | ···················· | |
| C | 984 hCoV-19/Italy/LOM-INMI-9214/2020\|EPI_ISL_460088\|20 | ···················· | ···················· | ···················· | ···················· | ···················· | ···················· | |
| C | 985 hCoV-19/Italy/LOM-INMI-9491/2020\|EPI_ISL_460089\|20 | ···················· | ···················· | ···················· | ···················· | ···················· | ···················· | |
| C | 986 hCoV-19/Italy/LOM-INMI-11550/2020\|EPI_ISL_460090\|2 | ···················· | ···················· | ···················· | ···················· | ···················· | ···················· | |
| C | 987 hCoV-19/Italy/LOM-INMI-5925/2020\|EPI_ISL_460091\|20 | ···················· | ···················· | ···················· | ···················· | ···················· | ···················· | |
| C | 988 hCoV-19/Italy/LOM-INMI-11951/2020\|EPI_ISL_460092\|2 | ···················· | ···················· | ···················· | ···················· | ···················· | ···················· | |
| C | 989 hCoV-19/Italy/LOM-INMI-12578/2020\|EPI_ISL_460093\|2 | ···················· | ···················· | ···················· | ···················· | ···················· | ···················· | |
| C | 990 hCoV-19/Italy/LOM-INMI-8058/2020\|EPI_ISL_460094\|20 | ···················· | ···················· | ···················· | ···················· | ···················· | ···················· | |
| C | 991 hCoV-19/Italy/LOM-INMI-9675/2020\|EPI_ISL_460095\|20 | ···················· | ···················· | ···················· | ···················· | ···················· | ···················· | |
| C | 992 hCoV-19/Russia/CRIE156278/2020\|EPI_ISL_460096\|2020 | ···················· | ···················· | ···················· | ···················· | ···················· | ···················· | |
| C | 993 hCoV-19/USA/MA-MGH-00568/2020\|EPI_ISL_460097\|2020- | ···················· | ···················· | ···················· | ···················· | ···················· | ···················· | |
| C | 994 hCoV-19/USA/MA-MGH-00277/2020\|EPI_ISL_460098\|2020- | ···················· | ···················· | ···················· | ···················· | ···················· | ···················· | |
| C | 995 hCoV-19/Netherlands/ZuidHolland_149/2020\|EPI_ISL_4 | ···················· | ···················· | ···················· | ···················· | ···················· | ···················· | |
| C | 996 hCoV-19/Netherlands/ZuidHolland_150/2020\|EPI_ISL_4 | ···················· | ···················· | ···················· | ···················· | ···················· | ···················· | |
| C | 997 hCoV-19/Netherlands/ZuidHolland_151/2020\|EPI_ISL_4 | ···················· | ···················· | ···················· | ···················· | ···················· | ···················· | |
| C | 998 hCoV-19/Netherlands/ZuidHolland_152/2020\|EPI_ISL_4 | ···················· | ···················· | ···················· | ···················· | ···················· | ···················· | |
| C | 999 hCoV-19/Netherlands/ZuidHolland_153/2020\|EPI_ISL_4 | ···················· | ···················· | ···················· | ···················· | ···················· | ···················· | |
| C | 1000 hCoV-19/Netherlands/ZuidHolland_159/2020\|EPI_ISL_4 | ···················· | ···················· | ···················· | ···················· | ···················· | ···················· | |
| C | 1001 hCoV-19/Netherlands/ZuidHolland_160/2020\|EPI_ISL_4 | ···················· | ···················· | ···················· | ···················· | ···················· | ···················· | |
| C | 1002 hCoV-19/Netherlands/ZuidHolland_161/2020\|EPI_ISL_4 | ···················· | ···················· | ···················· | ···················· | ···················· | ···················· | |
| C | 1003 hCoV-19/Netherlands/ZuidHolland_162/2020\|EPI_ISL_4 | ···················· | ···················· | ···················· | ···················· | ···················· | ···················· | |
| C | 1004 hCoV-19/Netherlands/ZuidHolland_163/2020\|EPI_ISL_4 | ···················· | ···················· | ···················· | ···················· | ···················· | ···················· | |
| C | 1005 hCoV-19/Netherlands/ZuidHolland_165/2020\|EPI_ISL_4 | ···················· | ···················· | ···················· | ···················· | ···················· | ···················· | |
| C | 1006 hCoV-19/Netherlands/ZuidHolland_166/2020\|EPI_ISL_4 | ···················· | ···················· | ···················· | ···················· | ···················· | ···················· | |
| C | 1007 hCoV-19/Netherlands/ZuidHolland_167/2020\|EPI_ISL_4 | ···················· | ···················· | ···················· | ···················· | ···················· | ···················· | |
| C | 1008 hCoV-19/Netherlands/ZuidHolland_168/2020\|EPI_ISL_4 | ···················· | ···················· | ···················· | ···················· | ···················· | ···················· | |
| C | 1009 hCoV-19/Netherlands/ZuidHolland_169/2020\|EPI_ISL_4 | ···················· | ···················· | ···················· | ···················· | ···················· | ···················· | |
| C | 1010 hCoV-19/Netherlands/ZuidHolland_171/2020\|EPI_ISL_4 | ···················· | ···················· | ···················· | ···················· | ···················· | ···················· | |
| C | 1011 hCoV-19/Netherlands/ZuidHolland_214/2020\|EPI_ISL_4 | ···················· | ···················· | ···················· | ···················· | ···················· | ···················· | |
| C | 1012 hCoV-19/Netherlands/ZuidHolland_215/2020\|EPI_ISL_4 | ···················· | ···················· | ···················· | ···················· | ···················· | ···················· | |
| C | 1013 hCoV-19/Netherlands/ZuidHolland_216/2020\|EPI_ISL_4 | ···················· | ···················· | ···················· | ···················· | ···················· | ···················· | |
| C | 1014 hCoV-19/Netherlands/ZuidHolland_217/2020\|EPI_ISL_4 | ···················· | ···················· | ···················· | ···················· | ···················· | ···················· | |
| C | 1015 hCoV-19/Netherlands/ZuidHolland_218/2020\|EPI_ISL_4 | ···················· | ···················· | ···················· | ···················· | ···················· | ···················· | |
| C | 1016 hCoV-19/Netherlands/ZuidHolland_219/2020\|EPI_ISL_4 | ···················· | ···················· | ···················· | ···················· | ···················· | ···················· | |
| C | 1017 hCoV-19/Netherlands/ZuidHolland_220/2020\|EPI_ISL_4 | ···················· | ···················· | ···················· | ···················· | ···················· | ···················· | |
| C | 1018 hCoV-19/Netherlands/ZuidHolland_221/2020\|EPI_ISL_4 | ···················· | ···················· | ···················· | ···················· | ···················· | ···················· | |
| C | 1019 hCoV-19/Netherlands/ZuidHolland_222/2020\|EPI_ISL_4 | ···················· | ···················· | ···················· | ···················· | ···················· | ···················· | |
| C | 1020 hCoV-19/Netherlands/ZuidHolland_223/2020\|EPI_ISL_4 | ···················· | ···················· | ···················· | ···················· | ···················· | ···················· | |
| C | 1021 hCoV-19/Netherlands/ZuidHolland_224/2020\|EPI_ISL_4 | ···················· | ···················· | ···················· | ···················· | ···················· | ···················· | |
| C | 1022 hCoV-19/Netherlands/ZuidHolland_225/2020\|EPI_ISL_4 | ···················· | ···················· | ···················· | ···················· | ···················· | ···················· | |
| C | 1023 hCoV-19/Netherlands/ZuidHolland_226/2020\|EPI_ISL_4 | ···················· | ···················· | ···················· | ···················· | ···················· | ···················· | |
| C | 1024 hCoV-19/USA/WA-UW-4657/2020\|EPI_ISL_461399\|2020-04 | ···················· | ···················· | ···················· | ···················· | ···················· | ···················· | |
| C | 1025 hCoV-19/England/NORW-E988D/2020\|EPI_ISL_448340\|202 | ···················· | ···················· | ···················· | ···················· | ···················· | ···················· | |
| C | 1026 hCoV-19/England/NORW-E98BA/2020\|EPI_ISL_448341\|202 | ··················T· | ···················· | ···················· | ···················· | ···················· | ···················· | 1 |
| C | 1027 hCoV-19/England/NORW-E98D8/2020\|EPI_ISL_448342\|202 | ···················· | ···················· | ···················· | ···················· | ···················· | ···················· | |
| C | 1028 hCoV-19/England/NORW-E994E/2020\|EPI_ISL_448344\|202 | ···················· | ···················· | ···················· | ···················· | ···················· | ···················· | |
| C | 1029 hCoV-19/England/NORW-E998A/2020\|EPI_ISL_448345\|202 | ···················· | ···················· | ···················· | ···················· | ···················· | ···················· | |
| C | 1030 hCoV-19/England/NORW-E99B7/2020\|EPI_ISL_448346\|202 | ···················· | ···················· | ···················· | ···················· | ···················· | ···················· | |
| C | 1031 hCoV-19/England/NORW-E9A0F/2020\|EPI_ISL_448348\|202 | ···················· | ···················· | ···················· | ···················· | ···················· | ···················· | |
| C | 1032 hCoV-19/England/NORW-E9AD2/2020\|EPI_ISL_448350\|202 | ···················· | ···················· | ···················· | ···················· | ···················· | ···················· | |

C 1033 hCoV-19/England/NORW-E9B1B/2020|EPI_ISL_448351|202 ···················· ···················· ···················· ···················· ···················· ····················
C 1034 hCoV-19/England/NORW-E9B48/2020|EPI_ISL_448352|202 ···················· ···················· ···················· ···················· ···················· ····················
C 1035 hCoV-19/England/NORW-E9C09/2020|EPI_ISL_448355|202 ···················· ···················· ···················· ···················· ···················· ····················
C 1036 hCoV-19/England/NORW-E9C54/2020|EPI_ISL_448357|202 ···················· ···················· ···················· ···················· ···················· ····················
C 1037 hCoV-19/England/NORW-E9C63/2020|EPI_ISL_448358|202 ···················· ···················· ···················· ···················· ···················· ····················
C 1038 hCoV-19/England/NORW-E9C72/2020|EPI_ISL_448359|202 ···················· ···················· ···················· ···················· ···················· ····················
C 1039 hCoV-19/England/LIVE-9D949/2020|EPI_ISL_449605|202 ···················· ·A·················· ···················· ···················· ···················· ····················
C 1040 hCoV-19/England/LIVE-9D9A3/2020|EPI_ISL_449608|202 ···················· ···················· ···················· ···················· ···················· ····················
C 1041 hCoV-19/England/LIVE-9DA37/2020|EPI_ISL_449611|202 ···················· ···················· ···················· ···················· ···················· ····················
C 1042 hCoV-19/England/LIVE-9DADD/2020|EPI_ISL_449614|202 ···················· ···················· ···················· ···················· ···················· ····················
C 1043 hCoV-19/England/LIVE-9DD10/2020|EPI_ISL_449623|202 ···················· ···················· ···················· ···················· ···················· ····················
C 1044 hCoV-19/England/LOND-D3B05/2020|EPI_ISL_449626|202 ···················· ···················· ···················· ···················· ···················· ····················
C 1045 hCoV-19/England/LOND-D4C7A/2020|EPI_ISL_449670|202 ···················· ···················· ···················· ···················· ···················· ····················
C 1046 hCoV-19/England/LOND-D4C98/2020|EPI_ISL_449672|202 ···················· ···················· ···················· ···················· ···················· ····················
C 1047 hCoV-19/England/LOND-D4CA7/2020|EPI_ISL_449673|202 ···················· ···················· ···················· ···················· ···················· ····················
C 1048 hCoV-19/England/LOND-D4CB6/2020|EPI_ISL_449674|202 ···················· ·A·················· ···················· ···················· ···················· ····················
C 1049 hCoV-19/England/LOND-D4CD4/2020|EPI_ISL_449676|202 ···················· ···················· ···················· ···················· ···················· ····················
C 1050 hCoV-19/England/LOND-D4CF2/2020|EPI_ISL_449678|202 ···················· ···················· ···················· ···················· ···················· ····················
C 1051 hCoV-19/England/LOND-D4D0E/2020|EPI_ISL_449679|202 ···················· ···················· ···················· ···················· ···················· ····················
C 1052 hCoV-19/England/LOND-D514D/2020|EPI_ISL_449680|202 ···················· ···················· ···················· ···················· ···················· ····················
C 1053 hCoV-19/England/LOND-D515C/2020|EPI_ISL_449681|202 ···················· ···················· ···················· ···················· ···················· ····················
C 1054 hCoV-19/England/LOND-D516B/2020|EPI_ISL_449682|202 ···················· ···················· ···················· ···················· ···················· ····················
C 1055 hCoV-19/England/LOND-D5189/2020|EPI_ISL_449683|202 ···················· ···················· ···················· ···················· ···················· ····················
C 1056 hCoV-19/England/LOND-D51C5/2020|EPI_ISL_449685|202 ···················· ···················· ···················· ···················· ···················· ····················
C 1057 hCoV-19/England/LOND-D51D4/2020|EPI_ISL_449686|202 ···················· ···················· ···················· ···················· ···················· ····················
C 1058 hCoV-19/England/LOND-D51E3/2020|EPI_ISL_449687|202 ···················· ···················· ···················· ···················· ···················· ····················
C 1059 hCoV-19/England/LOND-D51F2/2020|EPI_ISL_449688|202 ···················· ···················· ···················· ···················· ···················· ····················
C 1060 hCoV-19/England/LOND-D520E/2020|EPI_ISL_449689|202 ···················· ···················· ···················· ···················· ···················· ····················
C 1061 hCoV-19/USA/MI-MDHHS-SC20322/2020|EPI_ISL_450601|2 ···················· ···················· ···················· ···················· ···················· ····················
C 1062 hCoV-19/USA/MI-MDHHS-SC20325/2020|EPI_ISL_450604|2 ···················· ···················· ···················· ···················· ···················· ····················
C 1063 hCoV-19/USA/MI-MDHHS-SC20327/2020|EPI_ISL_450606|2 ···················· ···················· ···················· ···················· ···················· ····················
C 1064 hCoV-19/USA/MI-MDHHS-SC20328/2020|EPI_ISL_450607|2 ···················· ···················· ···················· ···················· ···················· ····················
C 1074 hCoV-19/Switzerland/VD-ETHZ-110508/2020|EPI_ISL_45 ···················· ···················· ···················· ···················· ···················· ····················
C 1077 hCoV-19/Croatia/ZG-297-20/2020|EPI_ISL_451934|2020 ···················· ···················· ···················· ···················· ···················· ····················
C 1078 hCoV-19/Belgium/20A30/2020|EPI_ISL_451935|2020-03- ···················· ···················· ···················· ···················· ···················· ····················
C 1079 hCoV-19/Germany/BY-MVP-0113/2020|EPI_ISL_451937|20 ···················· ···················· ···················· ···················· ···················· ····················
C 1080 hCoV-19/Poland/PL_P20/2020|EPI_ISL_451981|2020-04- ···················· ···················· ···················· ···················· ···················· ····················
C 1081 hCoV-19/Poland/PL_P21/2020|EPI_ISL_451982|2020-03- ···················· ···················· ···················· ···················· ···················· ····················
C 1082 hCoV-19/Poland/PL_P22/2020|EPI_ISL_451983|2020-04- ···················· ···················· ···················· ···················· ···················· ····················
C 1083 hCoV-19/Poland/PL_P23/2020|EPI_ISL_451984|2020-03- ···················· ···················· ···················· ···················· ···················· ····················
C 1084 hCoV-19/Poland/PL_P24/2020|EPI_ISL_451985|2020-03- ···················· ···················· ···················· ···················· ···················· ····················
C 1085 hCoV-19/Poland/PL_P25/2020|EPI_ISL_451986|2020-04- ···················· ···················· ···················· ···················· ···················· ····················
C 1086 hCoV-19/Poland/PL_P26/2020|EPI_ISL_451987|2020-04- ···················· ···················· ···················· ···················· ···················· ····················
C 1087 hCoV-19/Denmark/ALAB-HH-101/2020|EPI_ISL_451988|20 ···················· ···················· ···················· ···················· ···················· ····················
C 1088 hCoV-19/Denmark/ALAB-HH-102/2020|EPI_ISL_451989|20 ···················· ···················· ···················· ···················· ···················· ····················
C 1089 hCoV-19/Denmark/ALAB-HH-104/2020|EPI_ISL_451990|20 ···················· ···················· ···················· ···················· ···················· ····················
C 1090 hCoV-19/Denmark/ALAB-HH-106/2020|EPI_ISL_451991|20 ···················· ···················· ···················· ···················· ···················· ····················
C 1091 hCoV-19/Denmark/ALAB-HH-107/2020|EPI_ISL_451992|20 ···················· ···················· ···················· ···················· ···················· ····················
C 1092 hCoV-19/Denmark/ALAB-HH-108/2020|EPI_ISL_451993|20 ···················· ···················· ···················· ···················· ···················· ····················
C 1093 hCoV-19/Denmark/ALAB-HH-110/2020|EPI_ISL_451994|20 ···················· ···················· ···················· ···················· ···················· ····················
C 1094 hCoV-19/Denmark/ALAB-HH-111/2020|EPI_ISL_451995|20 ···················· ···················· ···················· ···················· ···················· ····················
C 1095 hCoV-19/Denmark/ALAB-HH-112/2020|EPI_ISL_451996|20 ···················· ···················· ···················· ···················· ···················· ····················
C 1096 hCoV-19/Denmark/ALAB-HH-113/2020|EPI_ISL_451997|20 ···················· ···················· ···················· ···················· ···················· ····················
C 1097 hCoV-19/Denmark/ALAB-HH-114/2020|EPI_ISL_451998|20 ···················· ···················· ···················· ···················· ···················· ····················
C 1098 hCoV-19/Denmark/ALAB-HH-115/2020|EPI_ISL_451999|20 ···················· ···················· ···················· ···················· ···················· ····················
C 1099 hCoV-19/Portugal/PT0413/2020|EPI_ISL_454137|2020-0 ···················· ···················· ···················· ···················· ···················· ····················
C 1100 hCoV-19/Portugal/PT0415/2020|EPI_ISL_454139|2020-0 ···················· ···················· ···················· ···················· ···················· ····················
C 1101 hCoV-19/Portugal/PT0417/2020|EPI_ISL_454141|2020-0 ···················· ···················· ···················· ···················· ···················· ····················
C 1102 hCoV-19/Portugal/PT0418/2020|EPI_ISL_454142|2020-0 ···················· ···················· ···················· ···················· ···················· ····················
C 1103 hCoV-19/Portugal/PT0419/2020|EPI_ISL_454143|2020-0 ···················· ···················· ···················· ···················· ···················· ····················
C 1104 hCoV-19/Portugal/PT0420/2020|EPI_ISL_454144|2020-0 ···················· ···················· ···················· ···················· ···················· ····················
C 1105 hCoV-19/Portugal/PT0421/2020|EPI_ISL_454145|2020-0 ···················· ···················· ···················· ···················· ···················· ····················
C 1106 hCoV-19/Portugal/PT0422/2020|EPI_ISL_454146|2020-0 ···················· ···················· ···················· ···················· ···················· ····················
C 1107 hCoV-19/Portugal/PT0423/2020|EPI_ISL_454147|2020-0 ···················· ···················· ···················· ···················· ···················· ····················
C 1108 hCoV-19/Portugal/PT0425/2020|EPI_ISL_454149|2020-0 ···················· ···················· ···················· ···················· ···················· ····················
C 1109 hCoV-19/Portugal/PT0426/2020|EPI_ISL_454150|2020-0 ···················· ···················· ···················· ···················· ···················· ····················
C 1110 hCoV-19/Portugal/PT0430/2020|EPI_ISL_454154|2020-0 ···················· ···················· ···················· ···················· ···················· ····················
C 1111 hCoV-19/Portugal/PT0434/2020|EPI_ISL_454158|2020-0 ···················· ···················· ···················· ···················· ···················· ····················
C 1112 hCoV-19/Portugal/PT0435/2020|EPI_ISL_454159|2020-0 ···················· ·A·················· ···················· ···················· ···················· ····················
C 1113 hCoV-19/Portugal/PT0438/2020|EPI_ISL_454162|2020-0 ···················· ···················· ···················· ···················· ···················· ····················
C 1114 hCoV-19/Portugal/PT0439/2020|EPI_ISL_454163|2020-0 ···················· ···················· ···················· ···················· ···················· ····················
C 1115 hCoV-19/Portugal/PT0443/2020|EPI_ISL_454167|2020-0 ···················· ·············A······ ···················· ···················· ···················· ····················
C 1116 hCoV-19/Portugal/PT0444/2020|EPI_ISL_454168|2020-0 ···················· ···················· ···················· ···················· ···················· ····················
C 1117 hCoV-19/Portugal/PT0445/2020|EPI_ISL_454169|2020-0 ···················· ···················· ···················· ···················· ···················· ····················
C 1118 hCoV-19/Portugal/PT0447/2020|EPI_ISL_454171|2020-0 ···················· ···················· ···················· ···················· ···················· ····················
C 1119 hCoV-19/Portugal/PT0448/2020|EPI_ISL_454172|2020-0 ···················· ···················· ···················· ···················· ···················· ····················
C 1120 hCoV-19/Portugal/PT0449/2020|EPI_ISL_454173|2020-0 ···················· ···················· ···················· ···················· ···················· ····················
C 1121 hCoV-19/Portugal/PT0450/2020|EPI_ISL_454174|2020-0 ···················· ···················· ···················· ···················· ···················· ····················

| | | | | | | | | |
|---|---|---|---|---|---|---|---|---|
| C | 1122 | hCoV-19/Portugal/PT0454/2020\|EPI_ISL_454178\|2020-0 | .................... | .................... | .................... | .................... | .................... | .................... |
| C | 1123 | hCoV-19/Portugal/PT0457/2020\|EPI_ISL_454181\|2020-0 | .................... | .................... | .................... | .................... | .................... | .................... |
| C | 1124 | hCoV-19/Portugal/PT0458/2020\|EPI_ISL_454182\|2020-0 | .................... | .................... | .................... | .................... | .................... | .................... |
| C | 1125 | hCoV-19/Portugal/PT0459/2020\|EPI_ISL_454183\|2020-0 | .................... | .................... | .................... | .................... | .................... | .................... |
| C | 1126 | hCoV-19/Portugal/PT0460/2020\|EPI_ISL_454184\|2020-0 | .................... | .................... | .................... | .................... | .................... | .................... |
| C | 1127 | hCoV-19/Portugal/PT0461/2020\|EPI_ISL_454185\|2020-0 | .................... | .................... | .................... | .................... | .................... | .................... |
| C | 1128 | hCoV-19/Portugal/PT0462/2020\|EPI_ISL_454186\|2020-0 | .................... | .................... | .................... | .................... | .................... | .................... |
| C | 1129 | hCoV-19/Portugal/PT0464/2020\|EPI_ISL_454188\|2020-0 | .................... | .................... | .................... | .................... | .................... | .................... |
| C | 1130 | hCoV-19/Portugal/PT0465/2020\|EPI_ISL_454189\|2020-0 | .................... | .................... | .................... | .................... | .................... | .................... |
| C | 1131 | hCoV-19/Portugal/PT0466/2020\|EPI_ISL_454190\|2020-0 | .................... | .................... | .................... | .................... | .................... | .................... |
| C | 1132 | hCoV-19/Portugal/PT0467/2020\|EPI_ISL_454191\|2020-0 | .................... | .................... | .................... | .................... | .................... | .................... |
| C | 1133 | hCoV-19/Portugal/PT0468/2020\|EPI_ISL_454192\|2020-0 | .................... | .................... | .................... | .................... | .................... | .................... |
| C | 1134 | hCoV-19/Portugal/PT0469/2020\|EPI_ISL_454193\|2020-0 | .................... | .................... | .................... | .................... | .................... | .................... |
| C | 1135 | hCoV-19/Portugal/PT0470a/2020\|EPI_ISL_454194\|2020- | .................... | .................... | .................... | .................... | .................... | .................... |
| C | 1136 | hCoV-19/Portugal/PT0471/2020\|EPI_ISL_454195\|2020-0 | .................... | .................... | .................... | .................... | .................... | .................... |
| C | 1137 | hCoV-19/Portugal/PT0472/2020\|EPI_ISL_454196\|2020-0 | .................... | .................... | .................... | .................... | .................... | .................... |
| C | 1138 | hCoV-19/Portugal/PT0473/2020\|EPI_ISL_454197\|2020-0 | .................... | .................... | .................... | .................... | .................... | .................... |
| C | 1139 | hCoV-19/Portugal/PT0474/2020\|EPI_ISL_454198\|2020-0 | .................... | .................... | .................... | .................... | .................... | .................... |
| C | 1140 | hCoV-19/Portugal/PT0475/2020\|EPI_ISL_454199\|2020-0 | .................... | .................... | .................... | .................... | .................... | .................... |
| C | 1141 | hCoV-19/Netherlands/NA_403/2020\|EPI_ISL_455205\|202 | .................... | .................... | .................... | .................... | .................... | .................... |
| C | 1142 | hCoV-19/Netherlands/NA_404/2020\|EPI_ISL_455206\|202 | .................... | .................... | .................... | .................... | .................... | .................... |
| C | 1143 | hCoV-19/Netherlands/NA_406/2020\|EPI_ISL_455208\|202 | .................... | .................... | .................... | .................... | .................... | .................... |
| C | 1144 | hCoV-19/Netherlands/NA_407/2020\|EPI_ISL_455209\|202 | .................... | .................... | .................... | .................... | .................... | .................... |
| C | 1145 | hCoV-19/Netherlands/NA_408/2020\|EPI_ISL_455210\|202 | .................... | .................... | .................... | .................... | .................... | .................... |
| C | 1146 | hCoV-19/Netherlands/NA_411/2020\|EPI_ISL_455213\|202 | .................... | .................... | .................... | .................... | .................... | .................... |
| C | 1147 | hCoV-19/Netherlands/NA_413/2020\|EPI_ISL_455215\|202 | .................... | .................... | .................... | .................... | .................... | .................... |
| C | 1148 | hCoV-19/Netherlands/NA_414/2020\|EPI_ISL_455216\|202 | .................... | .................... | .................... | .................... | .................... | .................... |
| C | 1149 | hCoV-19/Netherlands/NA_415/2020\|EPI_ISL_455217\|202 | .................... | .................... | .................... | .................... | .................... | .................... |
| C | 1150 | hCoV-19/Netherlands/NA_417/2020\|EPI_ISL_455219\|202 | .................... | .................... | .................... | .................... | .................... | .................... |
| C | 1151 | hCoV-19/Netherlands/NA_419/2020\|EPI_ISL_455221\|202 | .................... | .................... | .................... | .................... | .................... | .................... |
| C | 1152 | hCoV-19/Netherlands/NA_420/2020\|EPI_ISL_455222\|202 | .................... | .................... | .................... | .................... | .................... | .................... |
| C | 1153 | hCoV-19/Netherlands/NA_421/2020\|EPI_ISL_455223\|202 | .................... | .................... | .................... | .................... | .................... | .................... |
| C | 1154 | hCoV-19/Netherlands/NA_422/2020\|EPI_ISL_455224\|202 | .................... | .................... | .................... | .................... | .................... | .................... |
| C | 1155 | hCoV-19/Netherlands/NA_423/2020\|EPI_ISL_455225\|202 | .................... | .................... | .................... | .................... | .................... | .................... |
| C | 1156 | hCoV-19/Romania/284213/2020\|EPI_ISL_455468\|2020-05 | .................... | .................... | .................... | .................... | .................... | .................... |
| C | 1157 | hCoV-19/Romania/283584/2020\|EPI_ISL_455469\|2020-05 | .................... | .A.................. | .................... | .................... | .................... | .................... |
| C | 1158 | hCoV-19/Romania/284489/2020\|EPI_ISL_455470\|2020-05 | .................... | .................... | .................... | .................... | .................... | .................... |
| C | 1159 | hCoV-19/Romania/284508/2020\|EPI_ISL_455471\|2020-05 | .................... | .................... | .................... | .................... | .................... | .................... |
| C | 1160 | hCoV-19/Romania/284168/2020\|EPI_ISL_455472\|2020-05 | .................... | .A.................. | .................... | .................... | .................... | .................... |
| C | 1161 | hCoV-19/Romania/284056/2020\|EPI_ISL_455473\|2020-05 | .................... | .................... | .................... | .................... | .................... | .................... |
| C | 1162 | hCoV-19/Romania/284783/2020\|EPI_ISL_455474\|2020-05 | .................... | .................... | .................... | .................... | .................... | .................... |
| C | 1163 | hCoV-19/Romania/284762/2020\|EPI_ISL_455475\|2020-05 | .................... | .................... | .................... | .................... | .................... | .................... |
| C | 1164 | hCoV-19/Romania/285388/2020\|EPI_ISL_455476\|2020-05 | .................... | .................... | .................... | .................... | .................... | .................... |
| C | 1165 | hCoV-19/Romania/284371/2020\|EPI_ISL_455477\|2020-05 | .................... | .................... | .................... | .................... | .................... | .................... |
| C | 1166 | hCoV-19/India/OR-RMRC49/2020\|EPI_ISL_455478\|2020-0 | .................... | .................... | .................... | .................... | .................... | .................... |
| C | 1167 | hCoV-19/Serbia/CA3002-04/2020\|EPI_ISL_455480\|2020- | .................... | .................... | .................... | .................... | .................... | .................... |
| C | 1189 | hCoV-19/Scotland/CVR3369/2020\|EPI_ISL_456799\|2020- | .................... | .................... | .................... | .................... | .................... | .................... |
| C | 1190 | hCoV-19/USA/MD-HP00058/2020\|EPI_ISL_457800\|2020-03 | .................... | .................... | .................... | .................... | .................... | .................... |
| C | 1191 | hCoV-19/USA/DC-HP00059/2020\|EPI_ISL_457801\|2020-03 | .................... | .................... | .................... | .................... | .................... | .................... |
| C | 1193 | hCoV-19/USA/MD-HP00061/2020\|EPI_ISL_457803\|2020-03 | .................... | .................... | .................... | .................... | .................... | .................... |
| C | 1195 | hCoV-19/USA/DC-HP00063/2020\|EPI_ISL_457805\|2020-03 | .................... | .................... | .................... | .................... | .................... | .................... |
| C | 1196 | hCoV-19/USA/MD-HP00064/2020\|EPI_ISL_457806\|2020-03 | .................... | .................... | .................... | .................... | .................... | .................... |
| C | 1197 | hCoV-19/USA/MD-HP00065/2020\|EPI_ISL_457807\|2020-03 | .................... | .................... | .................... | .................... | .................... | .................... |
| C | 1198 | hCoV-19/USA/MD-HP00067/2020\|EPI_ISL_457808\|2020-03 | .................... | .................... | .................... | .................... | .................... | .................... |
| C | 1199 | hCoV-19/USA/DC-HP00116/2020\|EPI_ISL_457809\|2020-03 | .................... | .................... | .................... | .................... | .................... | .................... |
| C | 1200 | hCoV-19/USA/DC-HP00117/2020\|EPI_ISL_457810\|2020-03 | .................... | .................... | .................... | .................... | .................... | .................... |
| C | 1201 | hCoV-19/USA/MD-HP00118/2020\|EPI_ISL_457811\|2020-03 | .................... | .................... | .................... | .................... | .................... | .................... |
| C | 1202 | hCoV-19/USA/MD-HP00119/2020\|EPI_ISL_457812\|2020-03 | .................... | .................... | .................... | .................... | .................... | .................... |
| C | 1203 | hCoV-19/USA/MD-HP00120/2020\|EPI_ISL_457813\|2020-03 | .................... | .................... | .................... | .................... | .................... | .................... |
| C | 1204 | hCoV-19/USA/DC-HP00122/2020\|EPI_ISL_457814\|2020-03 | .................... | .................... | .................... | .................... | .................... | .................... |
| C | 1205 | hCoV-19/USA/MD-HP00127/2020\|EPI_ISL_457816\|2020-03 | .................... | .................... | .................... | .................... | .................... | .................... |
| C | 1206 | hCoV-19/USA/MD-HP00128/2020\|EPI_ISL_457817\|2020-03 | .................... | .................... | .................... | .................... | .................... | .................... |
| C | 1207 | hCoV-19/USA/MD-HP00129/2020\|EPI_ISL_457818\|2020-03 | .................... | .................... | .................... | .................... | .................... | .................... |
| C | 1208 | hCoV-19/USA/MD-HP00131/2020\|EPI_ISL_457819\|2020-03 | .................... | .................... | .................... | .................... | .................... | .................... |
| C | 1209 | hCoV-19/USA/MD-HP00132/2020\|EPI_ISL_457820\|2020-03 | .................... | .................... | .................... | .................... | .................... | .................... |
| C | 1210 | hCoV-19/USA/MD-HP00133/2020\|EPI_ISL_457821\|2020-03 | .................... | .................... | .................... | .................... | .................... | .................... |
| C | 1211 | hCoV-19/USA/DC-HP00135/2020\|EPI_ISL_457822\|2020-03 | .................... | .................... | .................... | .................... | .................... | .................... |
| C | 1212 | hCoV-19/USA/DC-HP00137/2020\|EPI_ISL_457823\|2020-03 | .................... | .................... | .................... | .................... | .................... | .................... |
| C | 1213 | hCoV-19/Turkey/Istanbul-BezmCoV1/2020\|EPI_ISL_4578 | .................... | .................... | .................... | .................... | .................... | .................... |
| C | 1214 | hCoV-19/Italy/SAR-ATS-AMC1-4634/2020\|EPI_ISL_45782 | .................... | .................... | .................... | .................... | .................... | .................... |
| C | 1215 | hCoV-19/Italy/LOM-ASST-AMC2-1245/2020\|EPI_ISL_4578 | .................... | .................... | .................... | .................... | .................... | .................... |
| C | 1216 | hCoV-19/Kenya/NIC_059/2020\|EPI_ISL_457827\|2020-03 | .................... | .................... | .................... | .................... | .................... | .................... |
| C | 1217 | hCoV-19/Kenya/NIC_084/2020\|EPI_ISL_457828\|2020-03 | .................... | .................... | .................... | .................... | .................... | .................... |
| C | 1219 | hCoV-19/Kenya/NIC_187/2020\|EPI_ISL_457833\|2020-03- | .................... | .................... | .................... | .................... | .................... | .................... |
| C | 1220 | hCoV-19/Kenya/NIC_569/2020\|EPI_ISL_457843\|2020-03- | .................... | .................... | .................... | .................... | .................... | .................... |
| C | 1221 | hCoV-19/Kenya/C1751/2020\|EPI_ISL_457867\|2020-04-22 | .................... | .................... | .................... | .................... | .................... | .................... |
| C | 1222 | hCoV-19/Kenya/C253/2020\|EPI_ISL_457868\|2020-04-03 | .................... | .................... | .................... | .................... | .................... | .................... |
| C | 1223 | hCoV-19/Kenya/C314/2020\|EPI_ISL_457875\|2020-04-04 | .................... | .................... | .................... | .................... | .................... | .................... |

| | | | | | | | | |
|---|---|---|---|---|---|---|---|---|
| C | 1224 | hCoV-19/Kenya/C2685/2020\|EPI_ISL_457884\|2020-04-25 | .................... | .................... | .................... | .................... | .................... | .................... |
| C | 1225 | hCoV-19/England/BRIS-1267A4/2020\|EPI_ISL_459001\|20 | .................... | .................... | .................... | .................... | .................... | .................... |
| C | 1226 | hCoV-19/England/BRIS-126917/2020\|EPI_ISL_459002\|20 | .................... | .................... | .................... | .................... | .................... | .................... |
| C | 1227 | hCoV-19/England/BRIS-126E36/2020\|EPI_ISL_459004\|20 | .................... | .................... | .................... | .................... | .................... | .................... |
| C | 1228 | hCoV-19/England/BRIS-1261A6/2020\|EPI_ISL_459005\|20 | .................... | .................... | .................... | .................... | .................... | .................... |
| C | 1229 | hCoV-19/England/BRIS-126610/2020\|EPI_ISL_459006\|20 | .................... | .................... | .................... | .................... | .................... | .................... |
| C | 1230 | hCoV-19/England/BRIS-126337/2020\|EPI_ISL_459007\|20 | .................... | .................... | .................... | .................... | .................... | .................... |
| C | 1231 | hCoV-19/England/BRIS-1260F4/2020\|EPI_ISL_459008\|20 | .................... | .................... | .................... | .................... | .................... | .................... |
| C | 1232 | hCoV-19/England/BRIS-126DDF/2020\|EPI_ISL_459010\|20 | .................... | .................... | .................... | .................... | .................... | .................... |
| C | 1233 | hCoV-19/England/BRIS-126B99/2020\|EPI_ISL_459012\|20 | .................... | .................... | .................... | .................... | .................... | .................... |
| C | 1234 | hCoV-19/England/BRIS-12680B/2020\|EPI_ISL_459017\|20 | .................... | .................... | .................... | .................... | .................... | .................... |
| C | 1235 | hCoV-19/Netherlands/NA_633/2020\|EPI_ISL_461068\|202 | .................... | .................... | .................... | .................... | .................... | .................... |
| C | 1236 | hCoV-19/Netherlands/NA_634/2020\|EPI_ISL_461069\|202 | .................... | .................... | .................... | .................... | .................... | .................... |
| C | 1237 | hCoV-19/Netherlands/NA_635/2020\|EPI_ISL_461070\|202 | .................... | .................... | .................... | .................... | .................... | .................... |
| C | 1238 | hCoV-19/Netherlands/NA_636/2020\|EPI_ISL_461071\|202 | .................... | .................... | .................... | .................... | .................... | .................... |
| C | 1239 | hCoV-19/Netherlands/NA_639/2020\|EPI_ISL_461074\|202 | .................... | .................... | .................... | .................... | .................... | .................... |
| C | 1240 | hCoV-19/Netherlands/NA_640/2020\|EPI_ISL_461075\|202 | .................... | .................... | .................... | .................... | .................... | .................... |
| C | 1241 | hCoV-19/Netherlands/NA_641/2020\|EPI_ISL_461076\|202 | .................... | .................... | .................... | .................... | .................... | .................... |
| C | 1242 | hCoV-19/Netherlands/NA_642/2020\|EPI_ISL_461077\|202 | .................... | .................... | .................... | .................... | .................... | .................... |
| C | 1243 | hCoV-19/Netherlands/NA_643/2020\|EPI_ISL_461078\|202 | .................... | .................... | .................... | .................... | .................... | .................... |
| C | 1244 | hCoV-19/Netherlands/NA_644/2020\|EPI_ISL_461079\|202 | .................... | .................... | .................... | .................... | .................... | .................... |
| C | 1245 | hCoV-19/Netherlands/NA_647/2020\|EPI_ISL_461082\|202 | .................... | .................... | .................... | .................... | .................... | .................... |
| C | 1246 | hCoV-19/Netherlands/NA_648/2020\|EPI_ISL_461083\|202 | .................... | .................... | .................... | .................... | .................... | .................... |
| C | 1247 | hCoV-19/Netherlands/NA_649/2020\|EPI_ISL_461084\|202 | .................... | .................... | .................... | .................... | .................... | .................... |
| C | 1248 | hCoV-19/Netherlands/NA_650/2020\|EPI_ISL_461085\|202 | .................... | .................... | .................... | .................... | .................... | .................... |
| C | 1249 | hCoV-19/Netherlands/NA_651/2020\|EPI_ISL_461086\|202 | .................... | .................... | .................... | .................... | .................... | .................... |
| C | 1250 | hCoV-19/Netherlands/NA_652/2020\|EPI_ISL_461087\|202 | .................... | .................... | .................... | .................... | .................... | .................... |
| C | 1251 | hCoV-19/Netherlands/NA_653/2020\|EPI_ISL_461088\|202 | .................... | .................... | .................... | .................... | .................... | .................... |
| C | 1252 | hCoV-19/Netherlands/NA_654/2020\|EPI_ISL_461089\|202 | .................... | .................... | .................... | .................... | .................... | .................... |
| C | 1253 | hCoV-19/Netherlands/NA_655/2020\|EPI_ISL_461090\|202 | .................... | .................... | .................... | .................... | .................... | .................... |
| C | 1254 | hCoV-19/Netherlands/NA_656/2020\|EPI_ISL_461091\|202 | .................... | .................... | .................... | .................... | .................... | .................... |
| C | 1255 | hCoV-19/Netherlands/NA_657/2020\|EPI_ISL_461092\|202 | .................... | .................... | .................... | .................... | .................... | .................... |
| C | 1256 | hCoV-19/Netherlands/NA_658/2020\|EPI_ISL_461093\|202 | .................... | .................... | .................... | .................... | .................... | .................... |
| C | 1257 | hCoV-19/Netherlands/NA_659/2020\|EPI_ISL_461094\|202 | .................... | .................... | .................... | .................... | .................... | .................... |
| C | 1258 | hCoV-19/Netherlands/NA_660/2020\|EPI_ISL_461095\|202 | .................... | .................... | .................... | .................... | .................... | .................... |
| C | 1259 | hCoV-19/Netherlands/NA_662/2020\|EPI_ISL_461097\|202 | .................... | .................... | .................... | .................... | .................... | .................... |
| C | 1260 | hCoV-19/Netherlands/NA_663/2020\|EPI_ISL_461098\|202 | .................... | .................... | .................... | .................... | .................... | .................... |
| C | 1261 | hCoV-19/Netherlands/ZuidHolland_127/2020\|EPI_ISL_4 | .................... | .................... | .................... | .................... | .................... | .................... |
| C | 1262 | hCoV-19/Netherlands/ZuidHolland_129/2020\|EPI_ISL_4 | .................... | .................... | .................... | .................... | .................... | .................... |
| C | 1263 | hCoV-19/Netherlands/ZuidHolland_130/2020\|EPI_ISL_4 | .................... | .................... | .................... | .................... | .................... | .................... |
| C | 1264 | hCoV-19/Netherlands/ZuidHolland_132/2020\|EPI_ISL_4 | .................... | .................... | .................... | .................... | .................... | .................... |
| C | 1265 | hCoV-19/Netherlands/ZuidHolland_134/2020\|EPI_ISL_4 | .................... | .................... | .................... | .................... | .................... | .................... |
| C | 1266 | hCoV-19/Netherlands/ZuidHolland_135/2020\|EPI_ISL_4 | .................... | .................... | .................... | .................... | .................... | .................... |
| C | 1267 | hCoV-19/Netherlands/ZuidHolland_136/2020\|EPI_ISL_4 | .................... | .................... | .................... | .................... | .................... | .................... |
| C | 1268 | hCoV-19/Netherlands/ZuidHolland_137/2020\|EPI_ISL_4 | .................... | .................... | .................... | .................... | .................... | .................... |
| C | 1269 | hCoV-19/Netherlands/ZuidHolland_138/2020\|EPI_ISL_4 | .................... | .................... | .................... | .................... | .................... | .................... |
| C | 1270 | hCoV-19/Netherlands/ZuidHolland_139/2020\|EPI_ISL_4 | .................... | .................... | .................... | .................... | .................... | .................... |
| C | 1271 | hCoV-19/Netherlands/ZuidHolland_140/2020\|EPI_ISL_4 | .................... | .................... | .................... | .................... | .................... | .................... |
| C | 1272 | hCoV-19/Netherlands/ZuidHolland_141/2020\|EPI_ISL_4 | .................... | .................... | .................... | .................... | .................... | .................... |
| C | 1273 | hCoV-19/Netherlands/ZuidHolland_142/2020\|EPI_ISL_4 | .................... | .................... | .................... | .................... | .................... | .................... |
| C | 1274 | hCoV-19/Netherlands/ZuidHolland_143/2020\|EPI_ISL_4 | .................... | .................... | .................... | .................... | .................... | .................... |
| C | 1275 | hCoV-19/Netherlands/ZuidHolland_144/2020\|EPI_ISL_4 | .................... | .................... | .................... | .................... | .................... | .................... |
| C | 1276 | hCoV-19/Netherlands/ZuidHolland_146/2020\|EPI_ISL_4 | .................... | .................... | .................... | .................... | .................... | .................... |
| C | 1277 | hCoV-19/Netherlands/ZuidHolland_147/2020\|EPI_ISL_4 | .................... | .................... | .................... | .................... | .................... | .................... |
| C | 1278 | hCoV-19/Netherlands/ZuidHolland_148/2020\|EPI_ISL_4 | .................... | .................... | .................... | .................... | .................... | .................... |
| C | 1279 | hCoV-19/Netherlands/ZuidHolland_155/2020\|EPI_ISL_4 | .................... | .................... | .................... | .................... | .................... | .................... |
| C | 1280 | hCoV-19/Netherlands/ZuidHolland_156/2020\|EPI_ISL_4 | .................... | .................... | .................... | .................... | .................... | .................... |
| C | 1281 | hCoV-19/England/CAMB-1AD7A5/2020\|EPI_ISL_448007\|20 | .................... | .................... | .................... | .................... | .................... | .................... |
| C | 1282 | hCoV-19/England/CAMB-1AD8DF/2020\|EPI_ISL_448021\|20 | .................... | .................... | .................... | .................... | .................... | .................... |
| C | 1283 | hCoV-19/England/CAMB-1AD8EE/2020\|EPI_ISL_448022\|20 | .................... | .................... | .................... | .................... | .................... | .................... |
| C | 1284 | hCoV-19/England/CAMB-1AD909/2020\|EPI_ISL_448023\|20 | .................... | .................... | .................... | .................... | .................... | .................... |
| C | 1285 | hCoV-19/England/CAMB-1AD918/2020\|EPI_ISL_448024\|20 | .................... | .................... | .................... | .................... | .................... | .................... |
| C | 1286 | hCoV-19/England/CAMB-1AD963/2020\|EPI_ISL_448028\|20 | .................... | .................... | .................... | .................... | .................... | .................... |
| C | 1289 | hCoV-19/England/CAMB-1ADAF7/2020\|EPI_ISL_448033\|20 | .................... | .................... | .................... | .................... | .................... | .................... |
| C | 1290 | hCoV-19/England/CAMB-1ADB12/2020\|EPI_ISL_448034\|20 | .................... | .................... | .................... | .................... | .................... | .................... |
| C | 1291 | hCoV-19/England/CAMB-1ADB21/2020\|EPI_ISL_448035\|20 | .................... | .................... | .................... | .................... | .................... | .................... |
| C | 1292 | hCoV-19/England/CAMB-1ADB30/2020\|EPI_ISL_448036\|20 | .................... | .................... | .................... | .................... | .................... | .................... |
| C | 1293 | hCoV-19/England/CAMB-1ADB5E/2020\|EPI_ISL_448037\|20 | .................... | .................... | .................... | .................... | .................... | .................... |
| C | 1294 | hCoV-19/England/CAMB-1ADB6D/2020\|EPI_ISL_448038\|20 | .................... | .................... | .................... | .................... | .................... | .................... |
| C | 1295 | hCoV-19/England/CAMB-1ADB7C/2020\|EPI_ISL_448039\|20 | .................... | .................... | .................... | .................... | .................... | .................... |
| C | 1296 | hCoV-19/England/CAMB-1ADB8B/2020\|EPI_ISL_448040\|20 | .................... | .................... | .................... | .................... | .................... | .................... |
| C | 1297 | hCoV-19/England/CAMB-1ADB9A/2020\|EPI_ISL_448041\|20 | .................... | .................... | .................... | .................... | .................... | .................... |
| C | 1298 | hCoV-19/England/CAMB-1ADBA9/2020\|EPI_ISL_448042\|20 | .................... | .................... | .................... | .................... | .................... | .................... |
| C | 1299 | hCoV-19/England/CAMB-1ADBB8/2020\|EPI_ISL_448043\|20 | .................... | .................... | .................... | .................... | .................... | .................... |
| C | 1300 | hCoV-19/England/CAMB-1ADBC7/2020\|EPI_ISL_448044\|20 | .................... | .................... | .................... | .................... | .................... | .................... |
| C | 1301 | hCoV-19/England/CAMB-1ADBE5/2020\|EPI_ISL_448046\|20 | .................... | .................... | .................... | .................... | .................... | .................... |
| C | 1302 | hCoV-19/England/CAMB-1ADBF4/2020\|EPI_ISL_448047\|20 | .................... | .................... | .................... | .................... | .................... | .................... |
| C | 1303 | hCoV-19/England/CAMB-1ADC79/2020\|EPI_ISL_448049\|20 | .................... | .................... | .................... | .................... | .................... | .................... |

| | | | | | | | | |
|---|---|---|---|---|---|---|---|---|
| C | 1304 hCoV-19/England/CAMB-1AE391/2020\|EPI_ISL_448051\|20 | ··················· | ··················· | ··················· | ··················· | ··················· | ··················· | |
| C | 1305 hCoV-19/England/CAMB-1AE3CE/2020\|EPI_ISL_448053\|20 | ··················· | ··················· | ··················· | ··················· | ··················· | ··················· | |
| C | 1306 hCoV-19/England/CAMB-1AEBB7/2020\|EPI_ISL_448056\|20 | ··················· | ··················· | ··················· | ··················· | ··················· | ··················· | |
| C | 1307 hCoV-19/England/CAMB-1AEBC6/2020\|EPI_ISL_448057\|20 | ··················· | ··················· | ··················· | ··················· | ··················· | ··················· | |
| C | 1308 hCoV-19/England/CAMB-1AEBE4/2020\|EPI_ISL_448059\|20 | ··················· | ··················· | ··················· | ··················· | ··················· | ··················· | |
| C | 1309 hCoV-19/England/CAMB-1AEBF3/2020\|EPI_ISL_448060\|20 | ··················· | ··················· | ··················· | ··················· | ··················· | ··················· | |
| C | 1310 hCoV-19/England/CAMB-1AEC2D/2020\|EPI_ISL_448063\|20 | ··················· | ··················· | ··················· | ··················· | ··················· | ··················· | |
| C | 1311 hCoV-19/England/CAMB-1AEC4B/2020\|EPI_ISL_448064\|20 | ··················· | ··················· | ··················· | ··················· | ··················· | ··················· | |
| C | 1312 hCoV-19/England/CAMB-1AEC5A/2020\|EPI_ISL_448065\|20 | ··················· | ··················· | ··················· | ··················· | ··················· | ··················· | |
| C | 1313 hCoV-19/England/CAMB-1AEC78/2020\|EPI_ISL_448066\|20 | ··················· | ··················· | ··················· | ··················· | ··················· | ··········A········ | |
| C | 1314 hCoV-19/England/CAMB-1AEC96/2020\|EPI_ISL_448068\|20 | ··················· | ··················· | ··················· | ··················· | ··················· | ··················· | |
| C | 1315 hCoV-19/England/CAMB-1AECE1/2020\|EPI_ISL_448072\|20 | ··················· | ··················· | ··················· | ··················· | ··················· | ··················· | |
| C | 1316 hCoV-19/England/CAMB-1AECF0/2020\|EPI_ISL_448073\|20 | ··················· | ··················· | ··················· | ··················· | ··················· | ··················· | |
| C | 1317 hCoV-19/England/CAMB-1AED39/2020\|EPI_ISL_448077\|20 | ··················· | ··················· | ··················· | ··················· | ··················· | ··················· | |
| C | 1318 hCoV-19/England/CAMB-1AED48/2020\|EPI_ISL_448078\|20 | ··················· | ··················· | ··················· | ··················· | ··················· | ··················· | |
| C | 1319 hCoV-19/England/CAMB-1AEF24/2020\|EPI_ISL_448079\|20 | ··················· | ··················· | ··················· | ··················· | ··················· | ··················· | |
| C | 1320 hCoV-19/England/CAMB-1AF55E/2020\|EPI_ISL_448080\|20 | ··················· | ··················· | ··················· | ··················· | ··················· | ··················· | |
| C | 1321 hCoV-19/England/CAMB-1AF56D/2020\|EPI_ISL_448081\|20 | ··················· | ··················· | ··················· | ··················· | ··················· | ··················· | |
| C | 1322 hCoV-19/England/CAMB-1AF58B/2020\|EPI_ISL_448082\|20 | ··················· | ··················· | ··················· | ··················· | ··················· | ··················· | |
| C | 1323 hCoV-19/England/CAMB-1AF5A9/2020\|EPI_ISL_448083\|20 | ··················· | ··················· | ··················· | ··················· | ··················· | ··················· | |
| C | 1324 hCoV-19/England/CAMB-1AF5C7/2020\|EPI_ISL_448084\|20 | ··················· | ··················· | ··················· | ··················· | ··················· | ··················· | |
| C | 1325 hCoV-19/England/CAMB-1AF5E5/2020\|EPI_ISL_448085\|20 | ··················· | ··················· | ··················· | ··················· | ··················· | ··················· | |
| C | 1326 hCoV-19/England/CAMB-1AF600/2020\|EPI_ISL_448086\|20 | ··················· | ·A················· | ··················· | ··················· | ··················· | ··················· | |
| C | 1327 hCoV-19/England/CAMB-1AF62E/2020\|EPI_ISL_448087\|20 | ··················· | ··················· | ··················· | ··················· | ··················· | ··················· | |
| C | 1328 hCoV-19/England/CAMB-1AF64C/2020\|EPI_ISL_448088\|20 | ··················· | ··················· | ··················· | ··················· | ··················· | ··················· | |
| C | 1329 hCoV-19/England/CAMB-1AF66A/2020\|EPI_ISL_448089\|20 | ··················· | ··················· | ··················· | ··················· | ··················· | ··················· | |
| C | 1330 hCoV-19/England/CAMB-1AF688/2020\|EPI_ISL_448090\|20 | ··················· | ·A················· | ··················· | ··················· | ··················· | ··················· | |
| C | 1332 hCoV-19/England/CAMB-1AFFAB/2020\|EPI_ISL_448092\|20 | ··················· | ··················· | ··················· | ··················· | ··················· | ··················· | |
| C | 1333 hCoV-19/England/CAMB-1AFFE7/2020\|EPI_ISL_448095\|20 | ··················· | ··················· | ··················· | ··················· | ··················· | ··················· | |
| C | 1334 hCoV-19/England/CAMB-1AFFF6/2020\|EPI_ISL_448096\|20 | ··················· | ··················· | ··················· | ··················· | ··················· | ··················· | |
| C | 1335 hCoV-19/England/CAMB-1B000E/2020\|EPI_ISL_448097\|20 | ··················· | ··················· | ··················· | ··················· | ······T············ | ··················· | |
| C | 1336 hCoV-19/England/CAMB-1B001D/2020\|EPI_ISL_448098\|20 | ··················· | ··················· | ··················· | ··················· | ··················· | ··················· | |
| C | 1337 hCoV-19/Scotland/EDB4367/2020\|EPI_ISL_449300\|2020- | ··················· | ··················· | ··················· | ··················· | ··················· | ··················· | |
| C | 1338 hCoV-19/Scotland/EDB4368/2020\|EPI_ISL_449301\|2020- | ··················· | ··················A | ··················· | ··················· | ··················· | ··················· | 1 |
| C | 1339 hCoV-19/Scotland/EDB4372/2020\|EPI_ISL_449302\|2020- | ··················· | ··················A | ··················· | ··················· | ··················· | ··················· | 1 |
| C | 1340 hCoV-19/Scotland/EDB4373/2020\|EPI_ISL_449303\|2020- | ··················· | ··················A | ··················· | ··················· | ··················· | ··················· | 1 |
| C | 1342 hCoV-19/India/MH-CCMB-NIV4/2020\|EPI_ISL_450324\|202 | ··················· | ··················· | ··················· | ··················· | ··················· | ··················· | |
| C | 1343 hCoV-19/India/MH-CCMB-NIV5/2020\|EPI_ISL_450325\|202 | ··················· | ··················· | ··················· | ··················· | ··················· | ··················· | |
| C | 1344 hCoV-19/India/TG-CCMB-J125/2020\|EPI_ISL_450326\|202 | ··················· | ··················· | ··················· | ··················· | ··················· | ··················· | |
| C | 1345 hCoV-19/India/TG-CCMB-J199/2020\|EPI_ISL_450327\|202 | ··················· | ··················· | ··················· | ··················· | ··················· | ··················· | |
| C | 1346 hCoV-19/India/TG-CCMB-J220/2020\|EPI_ISL_450331\|202 | ··················· | ··················· | ··················· | ··················· | ··················· | ··················· | |
| C | 1347 hCoV-19/India/TG-CCMB-J222/2020\|EPI_ISL_450332\|202 | ··················· | ··················· | ··················· | ··················· | ··················· | ··················· | |
| C | 1348 hCoV-19/Spain/CT-HUVH-VH7924/2020\|EPI_ISL_450337\|2 | ··················· | ··················· | ··················· | ··················· | ··················· | ··················· | |
| C | 1349 hCoV-19/Poland/IHG_PAS_1_77/2020\|EPI_ISL_450338\|20 | ··················· | ··················· | ··················· | ··················· | ··················· | ··················· | |
| C | 1350 hCoV-19/Bangladesh/BARJ-CVASU-CTG-501/2020\|EPI_ISL | ··················· | ··················· | ··················· | ··················· | ··················· | ··················· | |
| C | 1351 hCoV-19/Bangladesh/BARJ-CVASU-CTG-503/2020\|EPI_ISL | ··················· | ··················· | ··················· | ··················· | ··················· | ··················· | |
| C | 1352 hCoV-19/Bangladesh/BARJ-CVASU-CTG-511/2020\|EPI_ISL | ··················· | ·A················· | ··················· | ··················· | ··················· | ···A··············· | |
| C | 1353 hCoV-19/Bangladesh/BARJ-CVASU-CTG-517/2020\|EPI_ISL | ··················· | ·A················· | ··················· | ··················· | ··················· | ···A··············· | |
| C | 1354 hCoV-19/Bangladesh/BARJ-CVASU-CTG-518/2020\|EPI_ISL | ··················· | ··················· | ··················· | ··················· | ··················· | ··················· | |
| C | 1355 hCoV-19/Norway/Trondheim-S4/2020\|EPI_ISL_450346\|20 | ··················· | ··················· | ··················· | ··················· | ··················· | ··················· | |
| C | 1356 hCoV-19/Norway/Trondheim-S5/2020\|EPI_ISL_450347\|20 | ··················· | ··················· | ··················· | ··················· | ··················· | ··················· | |
| C | 1357 hCoV-19/Norway/Trondheim-S10/2020\|EPI_ISL_450348\|2 | ··················· | ··················· | ··················· | ··················· | ··················· | ··················· | |
| C | 1358 hCoV-19/Norway/Trondheim-S12/2020\|EPI_ISL_450349\|2 | ··················· | ··················· | ··················· | ··················· | ··················· | ··················· | |
| C | 1359 hCoV-19/Norway/Trondheim-S15/2020\|EPI_ISL_450350\|2 | ··················· | ··················· | ··················· | ··················· | ··················· | ··················· | |
| C | 1360 hCoV-19/Norway/Trondheim-E9/2020\|EPI_ISL_450351\|20 | ··················· | ··················· | ··················· | ··················· | ··················· | ··················· | |
| C | 1361 hCoV-19/Norway/Trondheim-E10/2020\|EPI_ISL_450352\|2 | ··················· | ··················· | ··················· | ··················· | ··················· | ··················· | |
| C | 1362 hCoV-19/USA/NY-NYUMC676/2020\|EPI_ISL_450393\|2020-0 | ··················· | ··················· | ··················· | ··················· | ··················· | ··················· | |
| C | 1363 hCoV-19/USA/NY-NYUMC677/2020\|EPI_ISL_450394\|2020-0 | ··················· | ··················· | ··················· | ··················· | ··················· | ··················· | |
| C | 1364 hCoV-19/USA/NY-NYUMC679/2020\|EPI_ISL_450396\|2020-0 | ··················· | ··················· | ··················· | ··················· | ··················· | ··················· | |
| C | 1365 hCoV-19/USA/NY-NYUMC680/2020\|EPI_ISL_450397\|2020-0 | ··················· | ··················· | ··················· | ··················· | ··················· | ··················· | |
| C | 1366 hCoV-19/USA/NY-NYUMC681/2020\|EPI_ISL_450398\|2020-0 | ··················· | ··················· | ··················· | ··················· | ··················· | ··················· | |
| C | 1367 hCoV-19/USA/NJ-NYUMC682/2020\|EPI_ISL_450399\|2020-0 | ··················· | ··················· | ··················· | ··················· | ··················· | ··················· | |
| C | 1368 hCoV-19/Australia/NSW338/2020\|EPI_ISL_451600\|2020- | ··················· | ··················· | ··················· | ··················· | ··················· | ··················· | |
| C | 1369 hCoV-19/Australia/NSW339/2020\|EPI_ISL_451601\|2020- | ··················· | ··················· | ··················· | ··················· | ··················· | ··················· | |
| C | 1370 hCoV-19/Australia/NSW340/2020\|EPI_ISL_451602\|2020- | ··················· | ··················· | ··················· | ··················· | ··················· | ··················· | |
| C | 1371 hCoV-19/Australia/NSW341/2020\|EPI_ISL_451603\|2020- | ··················· | ··················· | ··················· | ··················· | ··················· | ··················· | |
| C | 1372 hCoV-19/Australia/NSW342/2020\|EPI_ISL_451604\|2020- | ··················· | ··················· | ··················· | ··················· | ··················· | ··················· | |
| C | 1373 hCoV-19/Australia/NSW344/2020\|EPI_ISL_451605\|2020- | ··················· | ··················· | ··················· | ··················· | ··················· | ··················· | |
| C | 1374 hCoV-19/Australia/NSW345/2020\|EPI_ISL_451606\|2020- | ··················· | ··················· | ··················· | ··················· | ··················· | ··················· | |
| C | 1375 hCoV-19/Australia/NSW346/2020\|EPI_ISL_451607\|2020- | ··················· | ··················· | ··················· | ··················· | ··················· | ··················· | |
| C | 1376 hCoV-19/Australia/NSW347/2020\|EPI_ISL_451608\|2020- | ··················· | ··················· | ··················· | ··················· | ··················· | ··················· | |
| C | 1377 hCoV-19/Australia/NSW348/2020\|EPI_ISL_451609\|2020- | ··················· | ··················· | ··················· | ··················· | ··················· | ··················· | |
| C | 1378 hCoV-19/Australia/NSW349/2020\|EPI_ISL_451610\|2020- | ··················· | ··················· | ··················· | ··················· | ··················· | ··················· | |
| C | 1379 hCoV-19/Australia/NSW350/2020\|EPI_ISL_451611\|2020- | ··················· | ··················· | ··················· | ··················· | ··················· | ··················· | |
| C | 1380 hCoV-19/Australia/NSW351/2020\|EPI_ISL_451612\|2020- | ··················· | ··················· | ··················· | ··················· | ··················· | ··················· | |
| C | 1381 hCoV-19/Australia/NSW352/2020\|EPI_ISL_451613\|2020- | ··················· | ··················· | ··················· | ··················· | ··················· | ··················· | |
| C | 1382 hCoV-19/Australia/NSW353/2020\|EPI_ISL_451614\|2020- | ··················· | ··················· | ··················· | ··················· | ··················· | ··················· | |
| C | 1383 hCoV-19/Australia/NSW354/2020\|EPI_ISL_451615\|2020- | ··················· | ··················· | ··················· | ··················· | ··················· | ··················· | |

| | | | | | | | |
|---|---|---|---|---|---|---|---|
| C | 1384 hCoV-19/Australia/NSW355/2020\|EPI_ISL_451616\|2020- | ···················· | ···················· | ···················· | ···················· | ···················· | ···················· |
| C | 1385 hCoV-19/Australia/NSW356/2020\|EPI_ISL_451617\|2020- | ···················· | ···················· | ···················· | ···················· | ···················· | ···················· |
| C | 1386 hCoV-19/Australia/NSW357/2020\|EPI_ISL_451618\|2020- | ···················· | ···················· | ···················· | ···················· | ···················· | ···················· |
| C | 1387 hCoV-19/Australia/NSW358/2020\|EPI_ISL_451619\|2020- | ···················· | ···················· | ···················· | ···················· | ···················· | ···················· |
| C | 1388 hCoV-19/Poland/Sla5/2020\|EPI_ISL_451659\|2020-05-05 | ···················· | ···················· | ···················· | ···················· | ···················· | ···················· |
| C | 1389 hCoV-19/Poland/Sla8/2020\|EPI_ISL_451662\|2020-05-05 | ···················· | ···················· | ···················· | ···················· | ···················· | ···················· |
| C | 1390 hCoV-19/Poland/Sla9/2020\|EPI_ISL_451663\|2020-05-05 | ···················· | ···················· | ···················· | ···················· | ···················· | ···················· |
| C | 1391 hCoV-19/Poland/Sla10/2020\|EPI_ISL_451664\|2020-05-0 | ···················· | ···················· | ···················· | ···················· | ···················· | ···················· |
| C | 1392 hCoV-19/USA/NY-NYUMC751/2020\|EPI_ISL_451665\|2020-0 | ···················· | ···················· | ···················· | ···················· | ···················· | ···················· |
| C | 1393 hCoV-19/India/GJ-GBRC103/2020\|EPI_ISL_451666\|2020- | ···················· | ···················· | ···················· | ···················· | ···················· | ···················· |
| C | 1395 hCoV-19/Switzerland/VD-ETHZ-100076/2020\|EPI_ISL_45 | ···················· | ···················· | ···················· | ···················· | ···················· | ···················· |
| C | 1398 hCoV-19/Switzerland/BL-ETHZ-100081/2020\|EPI_ISL_45 | ···················· | ···················· | ···················· | ···················· | ···················· | ···················· |
| C | 1399 hCoV-19/Switzerland/ZH-ETHZ-100082/2020\|EPI_ISL_45 | ···················· | ···················· | ···················· | ···················· | ···················· | ···················· |
| C | 1400 hCoV-19/Switzerland/BS-ETHZ-100083/2020\|EPI_ISL_45 | ···················· | ···················· | ···················· | ···················· | ···················· | ···················· |
| C | 1401 hCoV-19/Switzerland/TI-ETHZ-100084/2020\|EPI_ISL_45 | ···················· | ···················· | ···················· | ···················· | ···················· | ···················· |
| C | 1402 hCoV-19/Switzerland/TI-ETHZ-100085/2020\|EPI_ISL_45 | ···················· | ···················· | ···················· | ···················· | ···················· | ···················· |
| C | 1403 hCoV-19/Switzerland/BL-ETHZ-100086/2020\|EPI_ISL_45 | ···················· | ···················· | ···················· | ···················· | ·······S············ | ···················· |
| C | 1406 hCoV-19/Switzerland/VS-ETHZ-100090/2020\|EPI_ISL_45 | ···················· | ···················· | ···················· | ···················· | ···················· | ···················· |
| C | 1407 hCoV-19/Switzerland/BL-ETHZ-100091/2020\|EPI_ISL_45 | ···················· | ···················· | ···················· | ···················· | ···················· | ···················· |
| C | 1408 hCoV-19/Switzerland/JU-ETHZ-100092/2020\|EPI_ISL_45 | ···················· | ···················· | ···················· | ···················· | ···················· | ···················· |
| C | 1409 hCoV-19/Switzerland/ZH-ETHZ-100093/2020\|EPI_ISL_45 | ···················· | ···················· | ···················· | ···················· | ···················· | ···················· |
| C | 1410 hCoV-19/Switzerland/SZ-ETHZ-100096/2020\|EPI_ISL_45 | ···················· | ···················· | ···················· | ···················· | ···················· | ···················· |
| C | 1411 hCoV-19/Switzerland/SZ-ETHZ-100097/2020\|EPI_ISL_45 | ···················· | ···················· | ···················· | ···················· | ···················· | ···················· |
| C | 1413 hCoV-19/Switzerland/VS-ETHZ-100099/2020\|EPI_ISL_45 | ···················· | ···················· | ···················· | ···················· | ···················· | ···················· |
| C | 1414 hCoV-19/Switzerland/BE-ETHZ-100108/2020\|EPI_ISL_45 | ···················· | ···················· | ···················· | ···················· | ···················· | ···················· |
| C | 1415 hCoV-19/Switzerland/BL-ETHZ-100109/2020\|EPI_ISL_45 | ···················· | ···················· | ···················· | ···················· | ···················· | ···················· |
| C | 1416 hCoV-19/Switzerland/AG-ETHZ-100110/2020\|EPI_ISL_45 | ···················· | ···················· | ···················· | ···················· | ···················· | ···················· |
| C | 1419 hCoV-19/Switzerland/BE-ETHZ-100113/2020\|EPI_ISL_45 | ···················· | ···················· | ···················· | ···················· | ···················· | ···················· |
| C | 1421 hCoV-19/Switzerland/BS-ETHZ-100115/2020\|EPI_ISL_45 | ···················· | ···················· | ···················· | ···················· | ···················· | ···················· |
| C | 1422 hCoV-19/England/CAMB-1B06DF/2020\|EPI_ISL_452910\|20 | ···················· | ···················· | ···················· | ···················· | ···················· | ···················· |
| C | 1423 hCoV-19/England/CAMB-1B06FD/2020\|EPI_ISL_452912\|20 | ···················· | ···················· | ···················· | ···················· | ···················· | ···················· |
| C | 1424 hCoV-19/England/CAMB-1B0709/2020\|EPI_ISL_452913\|20 | ···················· | ···················· | ···················· | ···················· | ···················· | ···················· |
| C | 1425 hCoV-19/England/CAMB-1B0718/2020\|EPI_ISL_452914\|20 | ···················· | ···················· | ···················· | ···················· | ···················· | ···················· |
| C | 1426 hCoV-19/England/CAMB-1B0727/2020\|EPI_ISL_452915\|20 | ···················· | ···················· | ···················· | ···················· | ···················· | ···················· |
| C | 1427 hCoV-19/England/CAMB-1B0736/2020\|EPI_ISL_452916\|20 | ···················· | ···················· | ···················· | ···················· | ···················· | ···················· |
| C | 1428 hCoV-19/England/CAMB-1B0745/2020\|EPI_ISL_452917\|20 | ···················· | ···················· | ···················· | ···················· | ···················· | ···················· |
| C | 1429 hCoV-19/England/CAMB-1B0754/2020\|EPI_ISL_452918\|20 | ···················· | ···················· | ···················· | ···················· | ···················· | ···················· |
| C | 1430 hCoV-19/England/CAMB-1B0781/2020\|EPI_ISL_452920\|20 | ···················· | ···················· | ···················· | ···················· | ···················· | ···················· |
| C | 1431 hCoV-19/England/CAMB-1B09D6/2020\|EPI_ISL_452926\|20 | ···················· | ···················· | ···················· | ···················· | ···················· | ···········A········ |
| C | 1432 hCoV-19/England/CAMB-1B0A00/2020\|EPI_ISL_452928\|20 | ···················· | ···················· | ···················· | ···················· | ···················· | ···················· |
| C | 1433 hCoV-19/England/CAMB-1B0A2E/2020\|EPI_ISL_452929\|20 | ···················· | ···················· | ···················· | ···················· | ···················· | ···················· |
| C | 1434 hCoV-19/England/CAMB-1B0A3D/2020\|EPI_ISL_452930\|20 | ···················· | ···················· | ···················· | ···················· | ···················· | ···················· |
| C | 1436 hCoV-19/England/CAMB-1B0A79/2020\|EPI_ISL_452934\|20 | ···················· | ···················· | ···················· | ···················· | ···················· | ···················· |
| C | 1437 hCoV-19/England/CAMB-1B0AA6/2020\|EPI_ISL_452936\|20 | ···················· | ···················· | ···················· | ···················· | ···················· | ···················· |
| C | 1438 hCoV-19/England/CAMB-1B0AB5/2020\|EPI_ISL_452937\|20 | ···················· | ···················· | ···················· | ···················· | ···················· | ···················· |
| C | 1439 hCoV-19/England/CAMB-1B0AF1/2020\|EPI_ISL_452938\|20 | ···················· | ···················· | ···················· | ···················· | ···················· | ···················· |
| C | 1440 hCoV-19/England/CAMB-1B0B1C/2020\|EPI_ISL_452940\|20 | ···················· | ···················· | ···················· | ···················· | ···················· | ···················· |
| C | 1441 hCoV-19/England/CAMB-1B0B3A/2020\|EPI_ISL_452942\|20 | ···················· | ···················· | ···················· | ···················· | ···················· | ···················· |
| C | 1442 hCoV-19/England/CAMB-1B0B58/2020\|EPI_ISL_452944\|20 | ···················· | ···················· | ···················· | ···················· | ···················· | ···················· |
| C | 1443 hCoV-19/England/CAMB-1B0B76/2020\|EPI_ISL_452946\|20 | ···················· | ···················· | ···················· | ···················· | ···················· | ···················· |
| C | 1444 hCoV-19/England/CAMB-1B0B94/2020\|EPI_ISL_452947\|20 | ···················· | ···················· | ···················· | ···················· | ···················· | ···················· |
| C | 1445 hCoV-19/England/CAMB-1B0BA3/2020\|EPI_ISL_452948\|20 | ···················· | ···················· | ···················· | ···················· | ···················· | ···················· |
| C | 1446 hCoV-19/England/CAMB-1B0BB2/2020\|EPI_ISL_452949\|20 | ···················· | ···················· | ···················· | ···················· | ···················· | ···················· |
| C | 1448 hCoV-19/England/CAMB-1B19E4/2020\|EPI_ISL_452994\|20 | ···················· | ···················· | ···················· | ···················· | ···················· | ···················· |
| C | 1450 hCoV-19/England/CAMB-1B1B1B/2020\|EPI_ISL_452998\|20 | ···················· | ···················· | ···················· | ···················· | ···················· | ···········A········ |
| C | 1451 hCoV-19/Netherlands/NA_388/2020\|EPI_ISL_455190\|202 | ···················· | ···················· | ···················· | ···················· | ···················· | ···················· |
| C | 1452 hCoV-19/Netherlands/NA_389/2020\|EPI_ISL_455191\|202 | ···················· | ···················· | ···················· | ···················· | ···················· | ···················· |
| C | 1453 hCoV-19/Netherlands/NA_390/2020\|EPI_ISL_455192\|202 | ···················· | ···················· | ···················· | ···················· | ···················· | ···················· |
| C | 1454 hCoV-19/Netherlands/NA_391/2020\|EPI_ISL_455193\|202 | ···················· | ···················· | ···················· | ···················· | ···················· | ···················· |
| C | 1455 hCoV-19/Netherlands/NA_393/2020\|EPI_ISL_455195\|202 | ···················· | ···················· | ···················· | ···················· | ···················· | ···················· |
| C | 1456 hCoV-19/Netherlands/NA_394/2020\|EPI_ISL_455196\|202 | ···················· | ···················· | ···················· | ···················· | ···················· | ···················· |
| C | 1457 hCoV-19/Netherlands/NA_396/2020\|EPI_ISL_455198\|202 | ···················· | ···················· | ···················· | ···················· | ···················· | ···················· |
| C | 1458 hCoV-19/Netherlands/NA_397/2020\|EPI_ISL_455199\|202 | ···················· | ···················· | ···················· | ···················· | ···················· | ···················· |
| C | 1459 hCoV-19/Netherlands/NA_398/2020\|EPI_ISL_455200\|202 | ···················· | ···················· | ···················· | ···················· | ···················· | ···················· |
| C | 1460 hCoV-19/Netherlands/NA_399/2020\|EPI_ISL_455201\|202 | ···················· | ···················· | ···················· | ···················· | ···················· | ···················· |
| C | 1461 hCoV-19/Netherlands/NA_400/2020\|EPI_ISL_455202\|202 | ···················· | ···················· | ···················· | ···················· | ···················· | ···················· |
| C | 1462 hCoV-19/Netherlands/NA_401/2020\|EPI_ISL_455203\|202 | ···················· | ···················· | ···················· | ···················· | ···················· | ···················· |
| C | 1463 hCoV-19/Netherlands/NA_409/2020\|EPI_ISL_455211\|202 | ···················· | ···················· | ···················· | ···················· | ···················· | ···················· |
| C | 1464 hCoV-19/Netherlands/NA_410/2020\|EPI_ISL_455212\|202 | ···················· | ···················· | ···················· | ···················· | ···················· | ···················· |
| C | 1465 hCoV-19/Netherlands/NA_412/2020\|EPI_ISL_455214\|202 | ···················· | ···················· | ···················· | ···················· | ···················· | ···················· |
| C | 1478 hCoV-19/Kenya/C1216/2020\|EPI_ISL_457854\|2020-04-19 | ···················· | ···················· | ···················· | ···················· | ···················· | ···················· |
| C | 1479 hCoV-19/Netherlands/Limburg_12/2020\|EPI_ISL_461000 | ···················· | ···················· | ···················· | ···················· | ···················· | ···················· |
| C | 1480 hCoV-19/Netherlands/Limburg_13/2020\|EPI_ISL_461001 | ···················· | ···················· | ···················· | ···················· | ···················· | ···················· |
| C | 1481 hCoV-19/Netherlands/Limburg_14/2020\|EPI_ISL_461002 | ···················· | ···················· | ···················· | ···················· | ···················· | ···················· |
| C | 1482 hCoV-19/Netherlands/Limburg_15/2020\|EPI_ISL_461003 | ···················· | ···················· | ···················· | ···················· | ···················· | ···················· |
| C | 1483 hCoV-19/Netherlands/Limburg_16/2020\|EPI_ISL_461004 | ···················· | ···················· | ···················· | ···················· | ···················· | ···················· |
| C | 1484 hCoV-19/Netherlands/Limburg_18/2020\|EPI_ISL_461006 | ···················· | ···················· | ···················· | ···················· | ···················· | ···················· |
| C | 1485 hCoV-19/Netherlands/Limburg_19/2020\|EPI_ISL_461007 | ···················· | ···················· | ···················· | ···················· | ···················· | ···················· |

```
C    1486 hCoV-19/Netherlands/Limburg_20/2020|EPI_ISL_461008   ····················  ····················  ····················  ····················  ····················  ····················
C    1487 hCoV-19/Netherlands/Limburg_21/2020|EPI_ISL_461009   ····················  ····················  ····················  ····················  ····················  ····················
C    1488 hCoV-19/Netherlands/Limburg_22/2020|EPI_ISL_461010   ····················  ····················  ····················  ····················  ····················  ····················
C    1489 hCoV-19/Netherlands/Limburg_23/2020|EPI_ISL_461011   ····················  ····················  ····················  ····················  ····················  ····················
C    1490 hCoV-19/Netherlands/Limburg_24/2020|EPI_ISL_461012   ····················  ····················  ····················  ····················  ····················  ····················
C    1491 hCoV-19/Netherlands/Limburg_25/2020|EPI_ISL_461013   ····················  ····················  ····················  ····················  ····················  ····················
C    1492 hCoV-19/Netherlands/Limburg_27/2020|EPI_ISL_461015   ····················  ····················  ····················  ····················  ····················  ····················
C    1493 hCoV-19/Netherlands/Limburg_28/2020|EPI_ISL_461016   ····················  ····················  ····················  ····················  ····················  ····················
C    1494 hCoV-19/Netherlands/Limburg_29/2020|EPI_ISL_461017   ····················  ····················  ····················  ····················  ····················  ····················
C    1495 hCoV-19/Netherlands/Limburg_30/2020|EPI_ISL_461018   ····················  ····················  ····················  ····················  ····················  ····················
C    1496 hCoV-19/Netherlands/Limburg_31/2020|EPI_ISL_461019   ····················  ····················  ····················  ····················  ····················  ····················
C    1497 hCoV-19/Netherlands/Limburg_32/2020|EPI_ISL_461020   ····················  ····················  ····················  ····················  ····················  ····················
C    1498 hCoV-19/Netherlands/Limburg_33/2020|EPI_ISL_461021   ····················  ····················  ····················  ····················  ····················  ····················
C    1499 hCoV-19/Netherlands/Limburg_34/2020|EPI_ISL_461022   ····················  ····················  ····················  ····················  ····················  ····················
C    1500 hCoV-19/Netherlands/Limburg_37/2020|EPI_ISL_461025   ····················  ····················  ····················  ····················  ····················  ····················
C    1501 hCoV-19/Netherlands/Limburg_38/2020|EPI_ISL_461026   ····················  ····················  ····················  ····················  ····················  ····················
C    1502 hCoV-19/Netherlands/Limburg_39/2020|EPI_ISL_461027   ····················  ····················  ····················  ····················  ····················  ····················
C    1503 hCoV-19/Netherlands/Limburg_41/2020|EPI_ISL_461029   ····················  ····················  ····················  ····················  ····················  ····················
C    1504 hCoV-19/Netherlands/Limburg_42/2020|EPI_ISL_461030   ····················  ····················  ····················  ····················  ····················  ····················
C    1505 hCoV-19/Netherlands/Limburg_43/2020|EPI_ISL_461031   ····················  ····················  ····················  ····················  ····················  ····················
C    1506 hCoV-19/Netherlands/Limburg_44/2020|EPI_ISL_461032   ····················  ····················  ····················  ····················  ····················  ····················
C    1507 hCoV-19/Netherlands/Limburg_45/2020|EPI_ISL_461033   ····················  ····················  ····················  ····················  ····················  ····················
C    1508 hCoV-19/Netherlands/Limburg_46/2020|EPI_ISL_461034   ····················  ····················  ····················  ····················  ····················  ····················
C    1509 hCoV-19/Netherlands/Limburg_47/2020|EPI_ISL_461035   ····················  ····················  ····················  ····················  ····················  ····················
C    1510 hCoV-19/Netherlands/Limburg_48/2020|EPI_ISL_461036   ····················  ····················  ····················  ····················  ····················  ····················
C    1511 hCoV-19/Netherlands/Limburg_49/2020|EPI_ISL_461037   ····················  ····················  ····················  ····················  ····················  ····················
C    1512 hCoV-19/Netherlands/Limburg_50/2020|EPI_ISL_461038   ····················  ····················  ····················  ····················  ····················  ····················
C    1513 hCoV-19/Netherlands/Limburg_51/2020|EPI_ISL_461039   ····················  ····················  ····················  ····················  ····················  ····················
C    1514 hCoV-19/Netherlands/Limburg_52/2020|EPI_ISL_461040   ····················  ····················  ····················  ····················  ····················  ····················
C    1515 hCoV-19/Netherlands/Limburg_53/2020|EPI_ISL_461041   ····················  ····················  ····················  ····················  ····················  ····················
C    1516 hCoV-19/Netherlands/Limburg_54/2020|EPI_ISL_461042   ····················  ····················  ····················  ····················  ····················  ····················
C    1517 hCoV-19/Netherlands/Limburg_55/2020|EPI_ISL_461043   ····················  ····················  ····················  ····················  ····················  ····················
C    1518 hCoV-19/Netherlands/Limburg_56/2020|EPI_ISL_461044   ····················  ····················  ····················  ····················  ····················  ····················
C    1519 hCoV-19/Netherlands/Limburg_59/2020|EPI_ISL_461045   ····················  ····················  ····················  ····················  ····················  ····················
C    1520 hCoV-19/Netherlands/Limburg_60/2020|EPI_ISL_461046   ····················  ····················  ····················  ····················  ····················  ····················
C    1521 hCoV-19/Netherlands/Limburg_61/2020|EPI_ISL_461047   ····················  ····················  ····················  ····················  ····················  ····················
C    1522 hCoV-19/Netherlands/Limburg_62/2020|EPI_ISL_461048   ····················  ····················  ····················  ····················  ····················  ····················
C    1523 hCoV-19/Netherlands/Limburg_66/2020|EPI_ISL_461051   ····················  ····················  ····················  ····················  ····················  ····················
C    1524 hCoV-19/Netherlands/Limburg_67/2020|EPI_ISL_461052   ····················  ····················  ····················  ····················  ····················  ····················
C    1525 hCoV-19/Netherlands/Limburg_68/2020|EPI_ISL_461053   ····················  ····················  ····················  ····················  ····················  ····················
C    1526 hCoV-19/Netherlands/Limburg_69/2020|EPI_ISL_461054   ····················  ····················  ····················  ····················  ····················  ····················
C    1527 hCoV-19/Netherlands/Limburg_70/2020|EPI_ISL_461055   ····················  ····················  ····················  ····················  ····················  ····················
C    1528 hCoV-19/Netherlands/Limburg_71/2020|EPI_ISL_461056   ····················  ····················  ····················  ····················  ····················  ····················
C    1529 hCoV-19/Netherlands/Limburg_72/2020|EPI_ISL_461057   ····················  ····················  ····················  ····················  ····················  ····················
C    1530 hCoV-19/Netherlands/Limburg_73/2020|EPI_ISL_461058   ····················  ····················  ····················  ····················  ····················  ····················
C    1531 hCoV-19/Netherlands/Limburg_74/2020|EPI_ISL_461059   ····················  ····················  ····················  ····················  ····················  ····················
C    1532 hCoV-19/Netherlands/Limburg_75/2020|EPI_ISL_461060   ····················  ····················  ····················  ····················  ····················  ····················
C    1533 hCoV-19/Netherlands/NA_628/2020|EPI_ISL_461063|202   ····················  ····················  ····················  ····················  ····················  ····················
C    1534 hCoV-19/Netherlands/NA_629/2020|EPI_ISL_461064|202   ····················  ····················  ····················  ····················  ····················  ····················
C    1535 hCoV-19/Netherlands/NA_630/2020|EPI_ISL_461065|202   ····················  ····················  ····················  ····················  ····················  ····················
C    1536 hCoV-19/Netherlands/NA_631/2020|EPI_ISL_461066|202   ····················  ····················  ····················  ····················  ····················  ····················
C    1537 hCoV-19/England/CAMB-1AD71E/2020|EPI_ISL_448000|20   ····················  ····················  ····················  ····················  ····················  ····················
C    1538 hCoV-19/England/CAMB-1AD72D/2020|EPI_ISL_448001|20   ····················  ····················  ····················  ····················  ····················  ····················
C    1539 hCoV-19/England/CAMB-1AD769/2020|EPI_ISL_448003|20   ····················  ····················  ····················  ····················  ····················  ····················
C    1540 hCoV-19/England/CAMB-1AD7B4/2020|EPI_ISL_448008|20   ····················  ····················  ····················  ····················  ····················  ····················
C    1541 hCoV-19/England/CAMB-1AD7D2/2020|EPI_ISL_448010|20   ····················  ····················  ····················  ····················  ····················  ····················
C    1542 hCoV-19/England/CAMB-1AD7E1/2020|EPI_ISL_448011|20   ····················  ····················  ····················  ····················  ····················  ····················
C    1543 hCoV-19/England/CAMB-1AD7F0/2020|EPI_ISL_448012|20   ····················  ····················  ····················  ····················  ····················  ····················
C    1544 hCoV-19/England/CAMB-1AD80C/2020|EPI_ISL_448013|20   ····················  ····················  ····················  ····················  ····················  ····················
C    1545 hCoV-19/England/CAMB-1AD81B/2020|EPI_ISL_448014|20   ····················  ····················  ····················  ····················  ····················  ····················
C    1546 hCoV-19/England/CAMB-1AD82A/2020|EPI_ISL_448015|20   ····················  ····················  ····················  ····················  ····················  ····················
C    1547 hCoV-19/England/CAMB-1AD857/2020|EPI_ISL_448016|20   ····················  ····················  ····················  ····················  ····················  ····················
C    1548 hCoV-19/England/CAMB-1AD866/2020|EPI_ISL_448017|20   ····················  ····················  ····················  ····················  ····················  ····················
C    1549 hCoV-19/England/CAMB-1AD893/2020|EPI_ISL_448018|20   ····················  ····················  ····················  ····················  ····················  ····················
C    1550 hCoV-19/England/CAMB-1AD8A2/2020|EPI_ISL_448019|20   ····················  ····················  ····················  ····················  ····················  ····················
C    1552 hCoV-19/Scotland/EDB4488/2020|EPI_ISL_449311|2020-   ····················  ····················  ····················  ····················  ····················  ····················
C    1553 hCoV-19/Scotland/EDB4492/2020|EPI_ISL_449312|2020-   ····················  ·A··················  ····················  ····················  ····················  ····················
C    1554 hCoV-19/Scotland/EDB4621/2020|EPI_ISL_449313|2020-   ····················  ····················  ····················  ····················  ····················  ····················
C    1555 hCoV-19/Scotland/EDB4654/2020|EPI_ISL_449315|2020-   ····················  ····················  ····················  ····················  ····················  ····················
C    1556 hCoV-19/Scotland/EDB4656/2020|EPI_ISL_449316|2020-   ····················  ····················  ····················  ····················  ····················  ····················
C    1557 hCoV-19/Scotland/EDB4661/2020|EPI_ISL_449318|2020-   ····················  ·A··················  ····················  ····················  ····················  ····················
C    1558 hCoV-19/Scotland/EDB4671/2020|EPI_ISL_449319|2020-   ····················  ·A··················  ····················  ····················  ····················  ····················
C    1559 hCoV-19/Scotland/EDB4679/2020|EPI_ISL_449320|2020-   ····················  ····················  ····················  ····················  ····················  ····················
C    1560 hCoV-19/Scotland/EDB4682/2020|EPI_ISL_449321|2020-   ····················  ·A··················  ····················  ····················  ····················  ····················
C    1561 hCoV-19/Scotland/EDB4705/2020|EPI_ISL_449322|2020-   ····················  ·A··················  ····················  ····················  ····················  ····················
C    1562 hCoV-19/Scotland/EDB4706/2020|EPI_ISL_449323|2020-   ····················  ·A··················  ····················  ····················  ····················  ····················
C    1563 hCoV-19/Scotland/EDB4708/2020|EPI_ISL_449324|2020-   ····················  ····················  ····················  ····················  ····················  ····················
C    1564 hCoV-19/Scotland/EDB4709/2020|EPI_ISL_449325|2020-   ····················  ····················  ····················  ····················  ····················  ····················
```

| | | | | | | | |
|---|---|---|---|---|---|---|---|
| C | 1565 hCoV-19/Scotland/EDB4710/2020\|EPI_ISL_449326\|2020- | ···················· | ···················· | ···················· | ···················· | ···················· | ···················· |
| C | 1566 hCoV-19/Scotland/EDB4711/2020\|EPI_ISL_449327\|2020- | ···················· | ···················· | ···················· | ···················· | ···················· | ···················· |
| C | 1567 hCoV-19/Scotland/EDB4712/2020\|EPI_ISL_449328\|2020- | ···················· | ···················· | ···················· | ···················· | ···················· | ···················· |
| C | 1568 hCoV-19/Scotland/EDB4713/2020\|EPI_ISL_449329\|2020- | ···················· | ···················· | ···················· | ···················· | ···················· | ···················· |
| C | 1569 hCoV-19/Canada/Qc-L00240556/2020\|EPI_ISL_450302\|20 | ···················· | ···················· | ···················· | ···················· | ···················· | ···················· |
| C | 1570 hCoV-19/Canada/Qc-L00240587/2020\|EPI_ISL_450303\|20 | ···················· | ···················· | ···················· | ···················· | ···················· | ···················· |
| C | 1571 hCoV-19/Canada/Qc-L00240652/2020\|EPI_ISL_450304\|20 | ···················· | ···················· | ···················· | ···················· | ···················· | ···················· |
| C | 1572 hCoV-19/Canada/Qc-L00240685/2020\|EPI_ISL_450305\|20 | ···················· | ···················· | ···················· | ···················· | ···················· | ···················· |
| C | 1573 hCoV-19/Canada/Qc-L00240857/2020\|EPI_ISL_450306\|20 | ···················· | ···················· | ···················· | ···················· | ···················· | ···················· |
| C | 1574 hCoV-19/Canada/Qc-L00240954/2020\|EPI_ISL_450307\|20 | ···················· | ···················· | ···················· | ···················· | ···················· | ···················· |
| C | 1575 hCoV-19/Canada/Qc-L00240979/2020\|EPI_ISL_450308\|20 | ···················· | ·A·················· | ···················· | ···················· | ···················· | ···················· |
| C | 1576 hCoV-19/Canada/Qc-L00241026/2020\|EPI_ISL_450309\|20 | ···················· | ···················· | ···················· | ···················· | ···················· | ···················· |
| C | 1577 hCoV-19/Canada/Qc-L00241042/2020\|EPI_ISL_450310\|20 | ···················· | ···················· | ···················· | ···················· | ···················· | ···················· |
| C | 1578 hCoV-19/Canada/Qc-L00241168/2020\|EPI_ISL_450311\|20 | ···················· | ···················· | ···················· | ···················· | ···················· | ···················· |
| C | 1579 hCoV-19/Canada/Qc-L00241192/2020\|EPI_ISL_450312\|20 | ···················· | ···················· | ···················· | ···················· | ···················· | ···················· |
| C | 1580 hCoV-19/Canada/Qc-L00241205/2020\|EPI_ISL_450313\|20 | ···················· | ···················· | ···················· | ···················· | ···················· | ···················· |
| C | 1581 hCoV-19/Canada/Qc-L00241208/2020\|EPI_ISL_450314\|20 | ···················· | ···················· | ···················· | ···················· | ···················· | ···················· |
| C | 1582 hCoV-19/Canada/Qc-L00241245/2020\|EPI_ISL_450315\|20 | ···················· | ·A·················· | ···················· | ···················· | ···················· | ···················· |
| C | 1583 hCoV-19/Canada/Qc-L00241292/2020\|EPI_ISL_450316\|20 | ···················· | ···················· | ···················· | ···················· | ···················· | ···················· |
| C | 1584 hCoV-19/Canada/Qc-L00241344/2020\|EPI_ISL_450317\|20 | ···················· | ···················· | ···················· | ···················· | ···················· | ···················· |
| C | 1585 hCoV-19/Canada/Qc-L00241355/2020\|EPI_ISL_450318\|20 | ···················· | ···················· | ···················· | ···················· | ···················· | ···················· |
| C | 1586 hCoV-19/India/MH-CCMB-NIV1/2020\|EPI_ISL_450321\|202 | ···················· | ···················· | ···················· | ···················· | ···················· | ···················· |
| C | 1587 hCoV-19/India/MH-CCMB-NIV2/2020\|EPI_ISL_450322\|202 | ···················· | ···················· | ···················· | ···················· | ···················· | ···················· |
| C | 1588 hCoV-19/India/MH-CCMB-NIV3/2020\|EPI_ISL_450323\|202 | ···················· | ···················· | ···················· | ···················· | ···················· | ···················· |
| C | 1589 hCoV-19/India/TG-CCMB-J283/2020\|EPI_ISL_450328\|202 | ···················· | ···················· | ···················· | ···················· | ···················· | ···················· |
| C | 1590 hCoV-19/India/TG-CCMB-J288/2020\|EPI_ISL_450329\|202 | ···················· | ···················· | ···················· | ···················· | ···················· | ···················· |
| C | 1591 hCoV-19/India/TG-CCMB-J375/2020\|EPI_ISL_450330\|202 | ···················· | ···················· | ···················· | ···················· | ···················· | ···················· |
| C | 1592 hCoV-19/Australia/NSW359/2020\|EPI_ISL_451620\|2020- | ···················· | ···················· | ···················· | ···················· | ···················· | ···················· |
| C | 1593 hCoV-19/Australia/NSW360/2020\|EPI_ISL_451621\|2020- | ···················· | ···················· | ···················· | ···················· | ···················· | ···················· |
| C | 1594 hCoV-19/Australia/NSW361/2020\|EPI_ISL_451622\|2020- | ···················· | ···················· | ···················· | ···················· | ···················· | ···················· |
| C | 1595 hCoV-19/Australia/NSW362/2020\|EPI_ISL_451623\|2020- | ···················· | ···················· | ···················· | ···················· | ···················· | ···················· |
| C | 1596 hCoV-19/Australia/NSW363/2020\|EPI_ISL_451624\|2020- | ···················· | ···················· | ···················· | ···················· | ···················· | ···················· |
| C | 1597 hCoV-19/Australia/NSW364/2020\|EPI_ISL_451625\|2020- | ···················· | ···················· | ···················· | ···················· | ···················· | ···················· |
| C | 1598 hCoV-19/Australia/NSW365/2020\|EPI_ISL_451626\|2020- | ···················· | ···················· | ···················· | ···················· | ···················· | ···················· |
| C | 1599 hCoV-19/Australia/NSW366/2020\|EPI_ISL_451627\|2020- | ···················· | ···················· | ···················· | ···················· | ···················· | ···················· |
| C | 1600 hCoV-19/Australia/NSW367/2020\|EPI_ISL_451628\|2020- | ···················· | ···················· | ···················· | ···················· | ···················· | ···················· |
| C | 1601 hCoV-19/Australia/NSW368/2020\|EPI_ISL_451629\|2020- | ···················· | ···················· | ···················· | ···················· | ···················· | ···················· |
| C | 1602 hCoV-19/Australia/NSW369/2020\|EPI_ISL_451630\|2020- | ···················· | ···················· | ···················· | ···················· | ···················· | ···················· |
| C | 1603 hCoV-19/Australia/NSW370/2020\|EPI_ISL_451631\|2020- | ···················· | ···················· | ···················· | ···················· | ···················· | ···················· |
| C | 1604 hCoV-19/Australia/NSW371/2020\|EPI_ISL_451632\|2020- | ···················· | ···················· | ···················· | ···················· | ···················· | ···················· |
| C | 1605 hCoV-19/Australia/NSW373/2020\|EPI_ISL_451633\|2020- | ···················· | ···················· | ···················· | ···················· | ···················· | ···················· |
| C | 1606 hCoV-19/Australia/NSW374/2020\|EPI_ISL_451634\|2020- | ···················· | ···················· | ···················· | ···················· | ···················· | ···················· |
| C | 1607 hCoV-19/Australia/NSW375/2020\|EPI_ISL_451635\|2020- | ···················· | ···················· | ···················· | ···················· | ···················· | ···················· |
| C | 1608 hCoV-19/Australia/NSW376/2020\|EPI_ISL_451636\|2020- | ···················· | ···················· | ···················· | ···················· | ···················· | ···················· |
| C | 1609 hCoV-19/Australia/NSW377/2020\|EPI_ISL_451637\|2020- | ···················· | ···················· | ···················· | ···················· | ···················· | ···················· |
| C | 1610 hCoV-19/Australia/NSW378/2020\|EPI_ISL_451638\|2020- | ···················· | ···················· | ···················· | ···················· | ···················· | ···················· |
| C | 1611 hCoV-19/Australia/NSW379/2020\|EPI_ISL_451639\|2020- | ···················· | ···················· | ···················· | ···················· | ···················· | ···················· |
| C | 1612 hCoV-19/Australia/NSW380/2020\|EPI_ISL_451640\|2020- | ···················· | ···················· | ···················· | ···················· | ···················· | ···················· |
| C | 1613 hCoV-19/Australia/NSW381/2020\|EPI_ISL_451641\|2020- | ···················· | ···················· | ···················· | ···················· | ···················· | ···················· |
| C | 1614 hCoV-19/Australia/NSW382/2020\|EPI_ISL_451642\|2020- | ···················· | ···················· | ···················· | ···················· | ···················· | ···················· |
| C | 1615 hCoV-19/Australia/NSW383/2020\|EPI_ISL_451643\|2020- | ···················· | ···················· | ···················· | ···················· | ···················· | ···················· |
| C | 1616 hCoV-19/Poland/Pom3/2020\|EPI_ISL_451644\|2020-05-02 | ···················· | ···················· | ···················· | ···················· | ···················· | ···················· |
| C | 1617 hCoV-19/Poland/Pom4/2020\|EPI_ISL_451645\|2020-05-02 | ···················· | ···················· | ···················· | ···················· | ···················· | ···················· |
| C | 1618 hCoV-19/Poland/Pom5/2020\|EPI_ISL_451646\|2020-04-22 | ···················· | ···················· | ···················· | ···················· | ···················· | ···················· |
| C | 1619 hCoV-19/Poland/Pom6/2020\|EPI_ISL_451647\|2020-04-23 | ···················· | ···················· | ···················· | ···················· | ···················· | ···················· |
| C | 1620 hCoV-19/Poland/Pom7/2020\|EPI_ISL_451648\|2020-04-26 | ···················· | ···················· | ···················· | ···················· | ···················· | ···················· |
| C | 1621 hCoV-19/Poland/Pom8/2020\|EPI_ISL_451649\|2020-04-27 | ···················· | ···················· | ···················· | ···················· | ···················· | ···················· |
| C | 1622 hCoV-19/Poland/Pom9/2020\|EPI_ISL_451650\|2020-04-27 | ···················· | ···················· | ···················· | ···················· | ···················· | ···················· |
| C | 1623 hCoV-19/Poland/Pom10/2020\|EPI_ISL_451651\|2020-04-2 | ···················· | ···················· | ···················· | ···················· | ···················· | ···················· |
| C | 1624 hCoV-19/Poland/Pom11/2020\|EPI_ISL_451652\|2020-04-3 | ···················· | ···················· | ···················· | ···················· | ···················· | ···················· |
| C | 1625 hCoV-19/Poland/Pom12/2020\|EPI_ISL_451653\|2020-05-0 | ···················· | ···················· | ···················· | ···················· | ···················· | ···················· |
| C | 1626 hCoV-19/Poland/Pom13/2020\|EPI_ISL_451654\|2020-05-0 | ···················· | ···················· | ···················· | ···················· | ···················· | ···················· |
| C | 1627 hCoV-19/Poland/Sla1/2020\|EPI_ISL_451655\|2020-05-05 | ···················· | ···················· | ···················· | ···················· | ···················· | ···················· |
| C | 1628 hCoV-19/Poland/Sla2/2020\|EPI_ISL_451656\|2020-05-05 | ···················· | ···················· | ···················· | ···················· | ···················· | ···················· |
| C | 1629 hCoV-19/Poland/Sla3/2020\|EPI_ISL_451657\|2020-05-05 | ···················· | ···················· | ···················· | ···················· | ···················· | ···················· |
| C | 1630 hCoV-19/Poland/Sla4/2020\|EPI_ISL_451658\|2020-05-05 | ···················· | ···················· | ···················· | ···················· | ···················· | ···················· |
| C | 1631 hCoV-19/Poland/Sla6/2020\|EPI_ISL_451660\|2020-05-05 | ···················· | ···················· | ···················· | ···················· | ···················· | ···················· |
| C | 1632 hCoV-19/Poland/Sla7/2020\|EPI_ISL_451661\|2020-05-05 | ···················· | ···················· | ···················· | ···················· | ···················· | ···················· |
| C | 1633 hCoV-19/England/CAMB-1B0675/2020\|EPI_ISL_452904\|20 | ···················· | ···················· | ···················· | ···················· | ···················· | ···················· |
| C | 1634 hCoV-19/England/CAMB-1B0693/2020\|EPI_ISL_452906\|20 | ···················· | ···················· | ···················· | ···················· | ···················· | ···················· |
| C | 1635 hCoV-19/England/CAMB-1B0BD0/2020\|EPI_ISL_452950\|20 | ···················· | ···················· | ···················· | ···················· | ···················· | ···················· |
| C | 1636 hCoV-19/England/CAMB-1B0C0A/2020\|EPI_ISL_452952\|20 | ···················· | ···················· | ···················· | ···················· | ···················· | ···················· |
| C | 1637 hCoV-19/England/CAMB-1B0C37/2020\|EPI_ISL_452953\|20 | ···················· | ···················· | ···················· | ···················· | ···················· | ··········N········· |
| C | 1638 hCoV-19/England/CAMB-1B0C46/2020\|EPI_ISL_452954\|20 | ···················· | ···················· | ···················· | ···················· | ···················· | ···················· |
| C | 1639 hCoV-19/England/CAMB-1B0C64/2020\|EPI_ISL_452955\|20 | ···················· | ···················· | ···················· | ···················· | ···················· | ···················· |
| C | 1640 hCoV-19/England/CAMB-1B0C73/2020\|EPI_ISL_452956\|20 | ···················· | ···················· | ···················· | ···················· | ···················· | ···················· |
| C | 1641 hCoV-19/England/CAMB-1B0C82/2020\|EPI_ISL_452957\|20 | ···················· | ···················· | ···················· | ···················· | ···················· | ···················· |
| C | 1642 hCoV-19/England/CAMB-1B0C91/2020\|EPI_ISL_452958\|20 | ···················· | ···················· | ···················· | ···················· | ···················· | ···················· |

| | | | | | | | | |
|---|---|---|---|---|---|---|---|---|
| C | 1643 | hCoV-19/England/CAMB-1B0CA0/2020\|EPI_ISL_452959\|20 | ···················· | ···················· | ···················· | ···················· | ···················· | ···················· |
| C | 1644 | hCoV-19/England/CAMB-1B0CBF/2020\|EPI_ISL_452960\|20 | ···················· | ···················· | ···················· | ···················· | ···················· | ···················· |
| C | 1645 | hCoV-19/England/CAMB-1B0CCE/2020\|EPI_ISL_452961\|20 | ···················· | ···················· | ···················· | ···················· | ···················· | ···················· |
| C | 1646 | hCoV-19/England/CAMB-1B0CDD/2020\|EPI_ISL_452962\|20 | ···················· | ···················· | ···················· | ···················· | ···················· | ···················· |
| C | 1647 | hCoV-19/England/CAMB-1B0CEC/2020\|EPI_ISL_452963\|20 | ···················· | ···················· | ···················· | ···················· | ···················· | ···················· |
| C | 1648 | hCoV-19/England/CAMB-1B0D16/2020\|EPI_ISL_452965\|20 | ···················· | ···················· | ···················· | ···················· | ···················· | ···················· |
| C | 1649 | hCoV-19/England/CAMB-1B0E5F/2020\|EPI_ISL_452968\|20 | ···················· | ···················· | ···················· | ···················· | ···················· | ···················· |
| C | 1650 | hCoV-19/England/CAMB-1B0E6E/2020\|EPI_ISL_452969\|20 | ···················· | ···················· | ···················· | ···················· | ···················· | ···················· |
| C | 1651 | hCoV-19/England/CAMB-1B0E8C/2020\|EPI_ISL_452971\|20 | ···················· | ···················· | ···················· | ···················· | ···················· | ···················· |
| C | 1652 | hCoV-19/England/CAMB-1B0E9B/2020\|EPI_ISL_452972\|20 | ···················· | ···················· | ···················· | ···················· | ···················· | ···················· |
| C | 1653 | hCoV-19/England/CAMB-1B0EAA/2020\|EPI_ISL_452973\|20 | ···················· | ···················· | ···················· | ···················· | ···················· | ···················· |
| C | 1654 | hCoV-19/England/CAMB-1B0EC8/2020\|EPI_ISL_452974\|20 | ···················· | ···················· | ···················· | ···················· | ···················· | ···················· |
| C | 1655 | hCoV-19/England/CAMB-1B0EF5/2020\|EPI_ISL_452977\|20 | ···················· | ···················· | ···················· | ···················· | ···················· | ···················· |
| C | 1656 | hCoV-19/England/CAMB-1B179F/2020\|EPI_ISL_452978\|20 | ···················· | ···················· | ···················· | ···················· | ···················· | ···················· |
| C | 1657 | hCoV-19/England/CAMB-1B17BD/2020\|EPI_ISL_452979\|20 | ···················· | ···················· | ···················· | ···················· | ···················· | ···················· |
| C | 1658 | hCoV-19/England/CAMB-1B17DB/2020\|EPI_ISL_452980\|20 | ···················· | ···················· | ···················· | ···················· | ···················· | ···················· |
| C | 1659 | hCoV-19/England/CAMB-1B1920/2020\|EPI_ISL_452982\|20 | ···················· | ···················· | ···················· | ···················· | ···················· | ···················· |
| C | 1660 | hCoV-19/England/CAMB-1B193F/2020\|EPI_ISL_452983\|20 | ···················· | ···················· | ···················· | ···················· | ···················· | ···················· |
| C | 1661 | hCoV-19/England/CAMB-1B194E/2020\|EPI_ISL_452984\|20 | ···················· | ···················· | ···················· | ···················· | ···················· | ···················· |
| C | 1662 | hCoV-19/England/CAMB-1B195D/2020\|EPI_ISL_452985\|20 | ···················· | ···················· | ···················· | ···················· | ···················· | ···················· |
| C | 1663 | hCoV-19/England/CAMB-1B196C/2020\|EPI_ISL_452986\|20 | ···················· | ···················· | ···················· | ···················· | ···················· | ···················· |
| C | 1665 | hCoV-19/Sweden/20-51317/2020\|EPI_ISL_455105\|2020-0 | ···················· | ···················· | ···················· | ···················· | ···················· | ···················· |
| C | 1666 | hCoV-19/Sweden/20-51349/2020\|EPI_ISL_455106\|2020-0 | ···················· | ···················· | ···················· | ···················· | ···················· | ···················· |
| C | 1667 | hCoV-19/Sweden/20-51387/2020\|EPI_ISL_455107\|2020-0 | ···················· | ···················· | ···················· | ···················· | ···················· | ···················· |
| C | 1668 | hCoV-19/Sweden/20-51391/2020\|EPI_ISL_455108\|2020-0 | ···················· | ···················· | ···················· | ···················· | ···················· | ···················· |
| C | 1669 | hCoV-19/Sweden/20-51426/2020\|EPI_ISL_455109\|2020-0 | ···················· | ···················· | ···················· | ···················· | ···················· | ···················· |
| C | 1670 | hCoV-19/Sweden/20-51445/2020\|EPI_ISL_455110\|2020-0 | ···················· | ···················· | ···················· | ···················· | ···················· | ···················· |
| C | 1671 | hCoV-19/Sweden/20-51474/2020\|EPI_ISL_455111\|2020-0 | ···················· | ···················· | ···················· | ···················· | ···················· | ···················· |
| C | 1672 | hCoV-19/Netherlands/Flevoland_3/2020\|EPI_ISL_45511 | ···················· | ···················· | ···················· | ···················· | ···················· | ···················· |
| C | 1673 | hCoV-19/Netherlands/Flevoland_4/2020\|EPI_ISL_45511 | ···················· | ···················· | ···················· | ···················· | ···················· | ···················· |
| C | 1674 | hCoV-19/Netherlands/Friesland_11/2020\|EPI_ISL_4551 | ···················· | ···················· | ···················· | ···················· | ···················· | ···················· |
| C | 1675 | hCoV-19/Netherlands/Friesland_12/2020\|EPI_ISL_4551 | ···················· | ···················· | ···················· | ···················· | ···················· | ···················· |
| C | 1676 | hCoV-19/Netherlands/Friesland_13/2020\|EPI_ISL_4551 | ···················· | ···················· | ···················· | ···················· | ···················· | ···················· |
| C | 1677 | hCoV-19/Netherlands/Friesland_14/2020\|EPI_ISL_4551 | ···················· | ···················· | ···················· | ···················· | ···················· | ···················· |
| C | 1678 | hCoV-19/Netherlands/Friesland_15/2020\|EPI_ISL_4551 | ···················· | ···················· | ···················· | ···················· | ···················· | ···················· |
| C | 1679 | hCoV-19/Netherlands/Friesland_16/2020\|EPI_ISL_4551 | ···················· | ···················· | ···················· | ···················· | ···················· | ···················· |
| C | 1680 | hCoV-19/Netherlands/Friesland_17/2020\|EPI_ISL_4551 | ···················· | ···················· | ···················· | ···················· | ···················· | ···················· |
| C | 1681 | hCoV-19/Netherlands/Friesland_18/2020\|EPI_ISL_4551 | ···················· | ···················· | ···················· | ···················· | ···················· | ···················· |
| C | 1682 | hCoV-19/Netherlands/Gelderland_13/2020\|EPI_ISL_455 | ···················· | ···················· | ···················· | ···················· | ···················· | ···················· |
| C | 1683 | hCoV-19/Netherlands/Gelderland_14/2020\|EPI_ISL_455 | ···················· | ···················· | ···················· | ···················· | ···················· | ···················· |
| C | 1684 | hCoV-19/Netherlands/NA_320/2020\|EPI_ISL_455125\|202 | ···················· | ···················· | ···················· | ···················· | ···················· | ···················· |
| C | 1685 | hCoV-19/Netherlands/NA_321/2020\|EPI_ISL_455126\|202 | ···················· | ···········A········ | ···················· | ···················· | ···················· | ···················· |
| C | 1686 | hCoV-19/Netherlands/NA_324/2020\|EPI_ISL_455129\|202 | ···················· | ···················· | ···················· | ···················· | ···················· | ···················· |
| C | 1687 | hCoV-19/Netherlands/NA_326/2020\|EPI_ISL_455131\|202 | ···················· | ···················· | ···················· | ···················· | ···················· | ···················· |
| C | 1688 | hCoV-19/Netherlands/NA_327/2020\|EPI_ISL_455132\|202 | ···················· | ···················· | ···················· | ···················· | ···················· | ···················· |
| C | 1689 | hCoV-19/Netherlands/NA_328/2020\|EPI_ISL_455133\|202 | ···················· | ···················· | ···················· | ···················· | ···················· | ···················· |
| C | 1690 | hCoV-19/Netherlands/NA_329/2020\|EPI_ISL_455134\|202 | ···················· | ···················· | ···················· | ···················· | ···················· | ···················· |
| C | 1691 | hCoV-19/Netherlands/NA_330/2020\|EPI_ISL_455135\|202 | ···················· | ···················· | ···················· | ···················· | ···················· | ···················· |
| C | 1692 | hCoV-19/Netherlands/NA_336/2020\|EPI_ISL_455141\|202 | ···················· | ···················· | ···················· | ···················· | ···················· | ···················· |
| C | 1693 | hCoV-19/Netherlands/NA_338/2020\|EPI_ISL_455143\|202 | ···················· | ···················· | ···················· | ···················· | ···················· | ···················· |
| C | 1694 | hCoV-19/Netherlands/NA_339/2020\|EPI_ISL_455144\|202 | ···················· | ···················· | ···················· | ···················· | ···················· | ···················· |
| C | 1695 | hCoV-19/Netherlands/NA_340/2020\|EPI_ISL_455145\|202 | ···················· | ···················· | ···················· | ···················· | ···················· | ···················· |
| C | 1696 | hCoV-19/Netherlands/NA_342/2020\|EPI_ISL_455147\|202 | ···················· | ···················· | ···················· | ···················· | ···················· | ···················· |
| C | 1697 | hCoV-19/Netherlands/NA_343/2020\|EPI_ISL_455148\|202 | ···················· | ···················· | ···················· | ···················· | ···················· | ···················· |
| C | 1698 | hCoV-19/Netherlands/NA_344/2020\|EPI_ISL_455149\|202 | ···················· | ···················· | ···················· | ···················· | ···················· | ···················· |
| C | 1699 | hCoV-19/Netherlands/NA_346/2020\|EPI_ISL_455151\|202 | ···················· | ···················· | ···················· | ···················· | ···················· | ···················· |
| C | 1700 | hCoV-19/Netherlands/NA_347/2020\|EPI_ISL_455152\|202 | ···················· | ···················· | ···················· | ···················· | ···················· | ···················· |
| C | 1701 | hCoV-19/Netherlands/NA_350/2020\|EPI_ISL_455155\|202 | ···················· | ···················· | ···················· | ···················· | ···················· | ···················· |
| C | 1702 | hCoV-19/Netherlands/NA_351/2020\|EPI_ISL_455156\|202 | ···················· | ···········A········ | ···················· | ···················· | ···················· | ···················· |
| C | 1703 | hCoV-19/Netherlands/NA_352/2020\|EPI_ISL_455157\|202 | ···················· | ···················· | ···················· | ···················· | ···················· | ···················· |
| C | 1704 | hCoV-19/Netherlands/NA_355/2020\|EPI_ISL_455159\|202 | ···················· | ···················· | ···················· | ···················· | ···················· | ···················· |
| C | 1705 | hCoV-19/Netherlands/NA_356/2020\|EPI_ISL_455160\|202 | ···················· | ···················· | ···················· | ···················· | ···················· | ···················· |
| C | 1706 | hCoV-19/Netherlands/NA_357/2020\|EPI_ISL_455161\|202 | ···················· | ···················· | ···················· | ···················· | ···················· | ···················· |
| C | 1707 | hCoV-19/Netherlands/NA_358/2020\|EPI_ISL_455162\|202 | ···················· | ···················· | ···················· | ···················· | ···················· | ···················· |
| C | 1708 | hCoV-19/Netherlands/NA_359/2020\|EPI_ISL_455163\|202 | ···················· | ···················· | ···················· | ···················· | ···················· | ···················· |
| C | 1709 | hCoV-19/Netherlands/NA_360/2020\|EPI_ISL_455164\|202 | ···················· | ···················· | ···················· | ···················· | ···················· | ···················· |
| C | 1710 | hCoV-19/Netherlands/NA_361/2020\|EPI_ISL_455165\|202 | ···················· | ···················· | ···················· | ···················· | ···················· | ···················· |
| C | 1711 | hCoV-19/Netherlands/NA_363/2020\|EPI_ISL_455167\|202 | ···················· | ···················· | ···················· | ···················· | ···················· | ···················· |
| C | 1712 | hCoV-19/Netherlands/NA_364/2020\|EPI_ISL_455168\|202 | ···················· | ···················· | ···················· | ···················· | ···················· | ···················· |
| C | 1713 | hCoV-19/Netherlands/NA_365/2020\|EPI_ISL_455169\|202 | ···················· | ···················· | ···················· | ···················· | ···················· | ···················· |
| C | 1714 | hCoV-19/Netherlands/NA_366/2020\|EPI_ISL_455170\|202 | ···················· | ···················· | ···················· | ···················· | ···················· | ···················· |
| C | 1715 | hCoV-19/Netherlands/NA_367/2020\|EPI_ISL_455171\|202 | ···················· | ·A·················· | ···················· | ···················· | ···················· | ···················· |
| C | 1716 | hCoV-19/Netherlands/NA_368/2020\|EPI_ISL_455172\|202 | ···················· | ···················· | ···················· | ···················· | ···················· | ···················· |
| C | 1717 | hCoV-19/Netherlands/NA_370/2020\|EPI_ISL_455173\|202 | ···················· | ···················· | ···················· | ···················· | ···················· | ···················· |
| C | 1718 | hCoV-19/Netherlands/NA_371/2020\|EPI_ISL_455174\|202 | ···················· | ···················· | ···················· | ···················· | ···················· | ···················· |
| C | 1719 | hCoV-19/Netherlands/NA_372/2020\|EPI_ISL_455175\|202 | ···················· | ···················· | ···················· | ···················· | ···················· | ···················· |
| C | 1720 | hCoV-19/Netherlands/NA_373/2020\|EPI_ISL_455176\|202 | ···················· | ···················· | ···················· | ···················· | ···················· | ···················· |
| C | 1721 | hCoV-19/Netherlands/NA_374/2020\|EPI_ISL_455177\|202 | ···················· | ···················· | ···················· | ···················· | ···················· | ···················· |

| | | | | | | | | |
|---|---|---|---|---|---|---|---|---|
| C | 1722 hCoV-19/Netherlands/NA_375/2020\|EPI_ISL_455178\|202 | ···················· | ···················· | ···················· | ···················· | ···················· | ···················· | |
| C | 1723 hCoV-19/Netherlands/NA_376/2020\|EPI_ISL_455179\|202 | ···················· | ···················· | ···················· | ···················· | ···················· | ···················· | |
| C | 1724 hCoV-19/Netherlands/NA_378/2020\|EPI_ISL_455180\|202 | ···················· | ···········A········ | ···················· | ···················· | ···················· | ···················· | |
| C | 1725 hCoV-19/Netherlands/NA_379/2020\|EPI_ISL_455181\|202 | ···················· | ···················· | ···················· | ···················· | ···················· | ···················· | |
| C | 1726 hCoV-19/Netherlands/NA_382/2020\|EPI_ISL_455184\|202 | ···················· | ···················· | ···················· | ···················· | ···················· | ···················· | |
| C | 1727 hCoV-19/Netherlands/NA_383/2020\|EPI_ISL_455185\|202 | ···················· | ···················· | ···················· | ···················· | ···················· | ···················· | |
| C | 1728 hCoV-19/Netherlands/NA_384/2020\|EPI_ISL_455186\|202 | ···················· | ···················· | ···················· | ···················· | ···················· | ···················· | |
| C | 1729 hCoV-19/Netherlands/NA_385/2020\|EPI_ISL_455187\|202 | ···················· | ···················· | ···················· | ···················· | ···················· | ···················· | |
| C | 1730 hCoV-19/Netherlands/NA_386/2020\|EPI_ISL_455188\|202 | ···················· | ···················· | ···················· | ···················· | ···················· | ···················· | |
| C | 1731 hCoV-19/Netherlands/NA_387/2020\|EPI_ISL_455189\|202 | ···················· | ···················· | ···················· | ···················· | ···················· | ···················· | |
| C | 1732 hCoV-19/Netherlands/ZuidHolland_111/2020\|EPI_ISL_4 | ···················· | ·A·················· | ···················· | ···················· | ···················· | ···················· | |
| C | 1733 hCoV-19/Netherlands/ZuidHolland_113/2020\|EPI_ISL_4 | ···················· | ·A·················· | ···················· | ···················· | ···················· | ···A················ | |
| C | 1738 hCoV-19/Australia/VIC1463/2020\|EPI_ISL_456445\|2020 | ···················· | ···················· | ···················· | ···················· | ···················· | ···················· | |
| C | 1746 hCoV-19/Australia/VIC1502/2020\|EPI_ISL_456462\|2020 | ···················· | ···················· | ···················· | ···················· | ···················· | ···················· | |
| C | 1747 hCoV-19/Australia/VIC1491/2020\|EPI_ISL_456463\|2020 | ···················· | ···················· | ···················· | ···················· | ···················· | ···················· | |
| C | 1755 hCoV-19/Australia/VIC1557/2020\|EPI_ISL_456479\|2020 | ···················· | ···················· | ···················· | ···················· | ···················· | ···················· | |
| C | 1767 hCoV-19/USA/MD-HP00113/2020\|EPI_ISL_457768\|2020-03 | ···················· | ···················· | ···················· | ···················· | ···················· | ···················· | |
| C | 1768 hCoV-19/USA/MD-HP00114/2020\|EPI_ISL_457769\|2020-03 | ···················· | ···················· | ···················· | ···················· | ···················· | ···················· | |
| C | 1772 hCoV-19/USA/MD-HP00154/2020\|EPI_ISL_457783\|2020-03 | ···················· | ···················· | ···················· | ···················· | ···················· | ···················· | |
| C | 1774 hCoV-19/USA/MD-HP00156/2020\|EPI_ISL_457785\|2020-03 | ···················· | ···················· | ···················· | ···················· | ···················· | ···················· | |
| C | 1775 hCoV-19/USA/MD-HP00158/2020\|EPI_ISL_457787\|2020-03 | ···················· | ···················· | ···················· | ···················· | ···················· | ···················· | |
| C | 1777 hCoV-19/USA/MD-HP00031/2020\|EPI_ISL_457789\|2020-03 | ···················· | ···················· | ···················· | ···················· | ···················· | ···················· | |
| C | 1778 hCoV-19/USA/MD-HP00017/2020\|EPI_ISL_457790\|2020-03 | ···················· | ···················· | ···················· | ···················· | ···················· | ···················· | |
| C | 1779 hCoV-19/USA/MD-HP00045/2020\|EPI_ISL_457791\|2020-03 | ···················· | ···················· | ···················· | ···················· | ···················· | ···················· | |
| C | 1780 hCoV-19/USA/MD-HP00048/2020\|EPI_ISL_457792\|2020-03 | ···················· | ···················· | ···················· | ···················· | ···················· | ···················· | |
| C | 1781 hCoV-19/USA/MD-HP00051/2020\|EPI_ISL_457795\|2020-03 | ···················· | ···················· | ···················· | ···················· | ···················· | ···················· | |
| C | 1782 hCoV-19/USA/DC-HP00052/2020\|EPI_ISL_457796\|2020-03 | ···················· | ···················· | ···················· | ···················· | ···················· | ···················· | |
| C | 1783 hCoV-19/USA/DC-HP00054/2020\|EPI_ISL_457797\|2020-03 | ···················· | ···················· | ···················· | ···················· | ···················· | ···················· | |
| C | 1784 hCoV-19/USA/MD-HP00055/2020\|EPI_ISL_457798\|2020-03 | ···················· | ···················· | ···················· | ···················· | ···················· | ···················· | |
| C | 1785 hCoV-19/USA/MD-HP00057/2020\|EPI_ISL_457799\|2020-03 | ···················· | ···················· | ···················· | ···················· | ···················· | ···················· | |
| C | 1786 hCoV-19/Uruguay/UY-NYUMC850/2020\|EPI_ISL_457946\|20 | ···················· | ···················· | ···················· | ···················· | ···················· | ···················· | |
| C | 1787 hCoV-19/Uruguay/UY-NYUMC860/2020\|EPI_ISL_457956\|20 | ···················· | ···················· | ···················· | ···················· | ···················· | ···················· | |
| C | 1788 hCoV-19/Uruguay/UY-NYUMC861/2020\|EPI_ISL_457957\|20 | ···················· | ···················· | ···················· | ···················· | ···················· | ···················· | |
| C | 1789 hCoV-19/Uruguay/UY-NYUMC862/2020\|EPI_ISL_457958\|20 | ···················· | ···················· | ···················· | ···················· | ···················· | ···················· | |
| C | 1790 hCoV-19/Uruguay/UY-NYUMC863/2020\|EPI_ISL_457959\|20 | ···················· | ···················· | ···················· | ···················· | ···················· | ···················· | |
| C | 1791 hCoV-19/Uruguay/UY-NYUMC865/2020\|EPI_ISL_457961\|20 | ···················· | ···················· | ···················· | ···················· | ···················· | ···················· | |
| C | 1792 hCoV-19/Uruguay/UY-NYUMC867/2020\|EPI_ISL_457963\|20 | ···················· | ···················· | ···················· | ···················· | ···················· | ···················· | |
| C | 1793 hCoV-19/Scotland/CVR1956/2020\|EPI_ISL_448140\|2020- | ···················· | ···················· | ···················· | ···················· | ···················· | ···················· | |
| C | 1794 hCoV-19/Scotland/CVR1958/2020\|EPI_ISL_448141\|2020- | ···················· | ···················· | ···················· | ···················· | ···················· | ···················· | |
| C | 1795 hCoV-19/Scotland/CVR2038/2020\|EPI_ISL_448142\|2020- | ···················· | ···················· | ···················· | ···················· | ···················· | ···················· | |
| C | 1796 hCoV-19/Scotland/CVR2052/2020\|EPI_ISL_448143\|2020- | ···················· | ···················· | ···················· | ···················· | ···················· | ···················· | |
| C | 1797 hCoV-19/Scotland/CVR2106/2020\|EPI_ISL_448144\|2020- | ···················· | ···················· | ···················· | ···················· | ···················· | ···················· | |
| C | 1798 hCoV-19/Scotland/CVR2110/2020\|EPI_ISL_448145\|2020- | ···················· | ···················· | ···················· | ···················· | ···················· | ···················· | |
| C | 1799 hCoV-19/Scotland/CVR2167/2020\|EPI_ISL_448147\|2020- | ···················· | ···················A | ···················· | ···················· | ···················· | ···················· | 1 |
| C | 1800 hCoV-19/Scotland/CVR2168/2020\|EPI_ISL_448148\|2020- | ···················· | ···················· | ···················· | ···················· | ···················· | ···················· | |
| C | 1801 hCoV-19/Scotland/CVR2169/2020\|EPI_ISL_448149\|2020- | ···················· | ···················A | ···················· | ···················· | ···················· | ···················· | 1 |
| C | 1802 hCoV-19/Scotland/CVR2173/2020\|EPI_ISL_448151\|2020- | ···················· | ·A·················· | ···················· | ···················· | ···················· | ···················· | |
| C | 1803 hCoV-19/Scotland/CVR2174/2020\|EPI_ISL_448152\|2020- | ···················· | ···················A | ···················· | ···················· | ···················· | ···················· | 1 |
| C | 1804 hCoV-19/Scotland/CVR2182/2020\|EPI_ISL_448153\|2020- | ···················· | ···················· | ···················· | ···················· | ···················· | ···················· | |
| C | 1805 hCoV-19/Scotland/CVR2193/2020\|EPI_ISL_448154\|2020- | ···················· | ···················· | ···················· | ···················· | ···················· | ···················· | |
| C | 1806 hCoV-19/Scotland/CVR2194/2020\|EPI_ISL_448155\|2020- | ···················· | ···················· | ···················· | ···················· | ···················· | ···················· | |
| C | 1807 hCoV-19/Scotland/CVR2195/2020\|EPI_ISL_448156\|2020- | ···················· | ···················· | ···················· | ···················· | ···················· | ···················· | |
| C | 1808 hCoV-19/Scotland/CVR2196/2020\|EPI_ISL_448157\|2020- | ···················· | ···················· | ···················· | ···················· | ···················· | ···················· | |
| C | 1809 hCoV-19/Scotland/CVR2198/2020\|EPI_ISL_448158\|2020- | ···················· | ···················· | ···················· | ···················· | ···················· | ···················· | |
| C | 1810 hCoV-19/Scotland/CVR2217/2020\|EPI_ISL_448161\|2020- | ···················· | ···················· | ···················· | ···················· | ···················· | ···················· | |
| C | 1811 hCoV-19/Scotland/CVR2218/2020\|EPI_ISL_448162\|2020- | ···················· | ···················· | ···················· | ···················· | ···················· | ···················· | |
| C | 1812 hCoV-19/Scotland/CVR2219/2020\|EPI_ISL_448163\|2020- | ···················· | ···················· | ···················· | ···················· | ···················· | ···················· | |
| C | 1813 hCoV-19/Scotland/CVR2221/2020\|EPI_ISL_448164\|2020- | ···················· | ···················A | ···················· | ···················· | ···················· | ···················· | 1 |
| C | 1814 hCoV-19/Scotland/CVR2222/2020\|EPI_ISL_448165\|2020- | ···················· | ···················· | ···················· | ···················· | ···················· | ···················· | |
| C | 1815 hCoV-19/Scotland/CVR2223/2020\|EPI_ISL_448166\|2020- | ···················· | ···················A | ···················· | ···················· | ···················· | ···················· | 1 |
| C | 1816 hCoV-19/Scotland/CVR2224/2020\|EPI_ISL_448167\|2020- | ···················· | ···················· | ···················· | ···················· | ···················· | ···················· | |
| C | 1817 hCoV-19/Scotland/CVR2226/2020\|EPI_ISL_448168\|2020- | ···················· | ···················· | ···················· | ···················· | ···················· | ···················· | |
| C | 1818 hCoV-19/Scotland/CVR2227/2020\|EPI_ISL_448169\|2020- | ···················· | ·A·················· | ···················· | ···················· | ···················· | ···················· | |
| C | 1819 hCoV-19/Scotland/CVR2252/2020\|EPI_ISL_448170\|2020- | ···················· | ···················· | ···················· | ···················· | ···················· | ···················· | |
| C | 1820 hCoV-19/Scotland/CVR2254/2020\|EPI_ISL_448171\|2020- | ···················· | ···················· | ···················· | ···················· | ···················· | ···················· | |
| C | 1821 hCoV-19/Scotland/CVR2278/2020\|EPI_ISL_448172\|2020- | ···················· | ···················· | ···················· | ···················· | ···················· | ···················· | |
| C | 1822 hCoV-19/Scotland/CVR2284/2020\|EPI_ISL_448175\|2020- | ···················· | ···················· | ···················· | ···················· | ···················· | ···················· | |
| C | 1823 hCoV-19/Scotland/CVR2294/2020\|EPI_ISL_448176\|2020- | ···················· | ···················· | ···················· | ···················· | ···················· | ···················· | |
| C | 1824 hCoV-19/Scotland/CVR2295/2020\|EPI_ISL_448177\|2020- | ···················· | ···················· | ···················· | ···················· | ···················· | ···················· | |
| C | 1825 hCoV-19/Scotland/CVR2316/2020\|EPI_ISL_448178\|2020- | ···················· | ···················· | ···················· | ···················· | ···················· | ···················· | |
| C | 1826 hCoV-19/Scotland/CVR2328/2020\|EPI_ISL_448179\|2020- | ···················· | ···················· | ···················· | ···················· | ···················· | ···················· | |
| C | 1827 hCoV-19/USA/CA-CZB-1203/2020\|EPI_ISL_450452\|2020-0 | ···················· | ···················· | ···················· | ···················· | ···················· | ···················· | |
| C | 1828 hCoV-19/USA/CA-CZB-1204/2020\|EPI_ISL_450453\|2020-0 | ···················· | ···················· | ···················· | ···················· | ···················· | ···················· | |
| C | 1829 hCoV-19/USA/CA-CZB-1205/2020\|EPI_ISL_450454\|2020-0 | ···················· | ···················· | ···················· | ···················· | ···················· | ···················· | |
| C | 1830 hCoV-19/USA/CA-CZB-1206/2020\|EPI_ISL_450455\|2020-0 | ···················· | ···················· | ···················· | ···················· | ···················· | ···················· | |
| C | 1831 hCoV-19/USA/CA-CZB-1207/2020\|EPI_ISL_450456\|2020-0 | ···················· | ···················· | ···················· | ···················· | ···················· | ···················· | |
| C | 1832 hCoV-19/USA/CA-CZB-1208/2020\|EPI_ISL_450457\|2020-0 | ···················· | ···················· | ···················· | ···················· | ···················· | ···················· | |
| C | 1833 hCoV-19/USA/CA-CZB-1209/2020\|EPI_ISL_450458\|2020-0 | ···················· | ···················· | ···················· | ···················· | ···················· | ···················· | |

| | | | | | | | |
|---|---|---|---|---|---|---|---|
| C | 1834 hCoV-19/USA/CA-CZB-1210/2020\|EPI_ISL_450459\|2020-0 | ···················· | ···················· | ···················· | ···················· | ···················· | ···················· |
| C | 1835 hCoV-19/USA/CA-CZB-1211/2020\|EPI_ISL_450460\|2020-0 | ···················· | ···················· | ···················· | ···················· | ···················· | ···················· |
| C | 1836 hCoV-19/USA/CA-CZB-1212/2020\|EPI_ISL_450461\|2020-0 | ···················· | ···················· | ···················· | ···················· | ···················· | ···················· |
| C | 1837 hCoV-19/USA/CA-CZB-1213/2020\|EPI_ISL_450462\|2020-0 | ···················· | ···················· | ···················· | ···················· | ···················· | ···················· |
| C | 1838 hCoV-19/USA/CA-CZB-1214/2020\|EPI_ISL_450463\|2020-0 | ···················· | ···················· | ···················· | ···················· | ···················· | ···················· |
| C | 1839 hCoV-19/USA/CA-CZB-1215/2020\|EPI_ISL_450464\|2020-0 | ···················· | ···················· | ···················· | ···················· | ···················· | ···················· |
| C | 1840 hCoV-19/USA/CA-CZB-1216/2020\|EPI_ISL_450465\|2020-0 | ···················· | ···················· | ···················· | ···················· | ···················· | ···················· |
| C | 1841 hCoV-19/USA/CA-CZB-1217/2020\|EPI_ISL_450466\|2020-0 | ···················· | ···················· | ···················· | ···················· | ···················· | ···················· |
| C | 1842 hCoV-19/USA/CA-CZB-1218/2020\|EPI_ISL_450467\|2020-0 | ···················· | ···················· | ···················· | ···················· | ···················· | ···················· |
| C | 1843 hCoV-19/USA/CA-CZB-1219/2020\|EPI_ISL_450468\|2020-0 | ···················· | ···················· | ···················· | ···················· | ···················· | ···················· |
| C | 1844 hCoV-19/USA/CA-CZB-1220/2020\|EPI_ISL_450469\|2020-0 | ···················· | ···················· | ···················· | ···················· | ···················· | ···················· |
| C | 1845 hCoV-19/USA/CA-CZB-1221/2020\|EPI_ISL_450470\|2020-0 | ···················· | ···················· | ···················· | ···················· | ···················· | ···················· |
| C | 1846 hCoV-19/USA/CA-CZB-1222/2020\|EPI_ISL_450471\|2020-0 | ···················· | ···················· | ···················· | ···················· | ···················· | ···················· |
| C | 1847 hCoV-19/USA/CA-CZB-1223/2020\|EPI_ISL_450472\|2020-0 | ···················· | ···················· | ···················· | ···················· | ···················· | ···················· |
| C | 1848 hCoV-19/USA/CA-CZB-1224/2020\|EPI_ISL_450473\|2020-0 | ···················· | ···················· | ···················· | ···················· | ···················· | ···················· |
| C | 1849 hCoV-19/USA/CA-CZB-1225/2020\|EPI_ISL_450474\|2020-0 | ···················· | ···················· | ···················· | ···················· | ···················· | ···················· |
| C | 1850 hCoV-19/USA/CA-CZB-1226/2020\|EPI_ISL_450475\|2020-0 | ···················· | ···················· | ···················· | ···················· | ···················· | ···················· |
| C | 1851 hCoV-19/USA/CA-CZB-1227/2020\|EPI_ISL_450476\|2020-0 | ···················· | ···················· | ···················· | ···················· | ···················· | ···················· |
| C | 1852 hCoV-19/USA/CA-CZB-1228/2020\|EPI_ISL_450477\|2020-0 | ···················· | ···················· | ···················· | ···················· | ···················· | ···················· |
| C | 1853 hCoV-19/USA/CA-CZB-1229/2020\|EPI_ISL_450478\|2020-0 | ···················· | ···················· | ···················· | ···················· | ···················· | ···················· |
| C | 1854 hCoV-19/USA/CA-CZB-1230/2020\|EPI_ISL_450479\|2020-0 | ···················· | ···················· | ···················· | ···················· | ···················· | ···················· |
| C | 1855 hCoV-19/USA/CA-CZB-1231/2020\|EPI_ISL_450480\|2020-0 | ···················· | ···················· | ···················· | ···················· | ···················· | ···················· |
| C | 1856 hCoV-19/USA/CA-CZB-1232/2020\|EPI_ISL_450481\|2020-0 | ···················· | ···················· | ···················· | ···················· | ···················· | ···················· |
| C | 1857 hCoV-19/Switzerland/VD-ETHZ-100836/2020\|EPI_ISL_45 | ···················· | ···················· | ···················· | ···················· | ···················· | ···················· |
| C | 1860 hCoV-19/Switzerland/VD-ETHZ-100839/2020\|EPI_ISL_45 | ···················· | ···················· | ···················· | ···················· | ···················· | ···················· |
| C | 1861 hCoV-19/Switzerland/GR-ETHZ-100840/2020\|EPI_ISL_45 | ···················· | ···················· | ···················· | ···················· | ···················· | ···················· |
| C | 1864 hCoV-19/Switzerland/GR-ETHZ-100844/2020\|EPI_ISL_45 | ···················· | ···················· | ···················· | ···················· | ···················· | ···················· |
| C | 1868 hCoV-19/Switzerland/BE-ETHZ-101129/2020\|EPI_ISL_45 | ···················· | ···················· | ···················· | ···················· | ···················· | ···················· |
| C | 1869 hCoV-19/Switzerland/ZH-ETHZ-101130/2020\|EPI_ISL_45 | ···················· | ···················· | ···················· | ···················· | ···················· | ···················· |
| C | 1870 hCoV-19/Switzerland/ZH-ETHZ-101131/2020\|EPI_ISL_45 | ···················· | ···················· | ···················· | ···················· | ···················· | ···················· |
| C | 1871 hCoV-19/Switzerland/ZH-ETHZ-101132/2020\|EPI_ISL_45 | ···················· | ···················· | ···················· | ···················· | ·······S············ | ···················· |
| C | 1872 hCoV-19/Portugal/PT0285/2020\|EPI_ISL_454000\|2020-0 | ···················· | ···················· | ···················· | ···················· | ···················· | ···················· |
| C | 1873 hCoV-19/Portugal/PT0286a/2020\|EPI_ISL_454001\|2020- | ···················· | ···················· | ···················· | ···················· | ···················· | ···················· |
| C | 1874 hCoV-19/Portugal/PT0287/2020\|EPI_ISL_454003\|2020-0 | ···················· | ···················· | ···················· | ···················· | ···················· | ···················· |
| C | 1875 hCoV-19/Portugal/PT0288/2020\|EPI_ISL_454004\|2020-0 | ···················· | ···················· | ···················· | ···················· | ···················· | ···················· |
| C | 1876 hCoV-19/Portugal/PT0289/2020\|EPI_ISL_454005\|2020-0 | ···················· | ···················· | ···················· | ···················· | ···················· | ···················· |
| C | 1877 hCoV-19/Portugal/PT0290/2020\|EPI_ISL_454006\|2020-0 | ···················· | ···················· | ···················· | ···················· | ···················· | ···················· |
| C | 1878 hCoV-19/Portugal/PT0291/2020\|EPI_ISL_454007\|2020-0 | ···················· | ···················· | ···················· | ···················· | ···················· | ···················· |
| C | 1879 hCoV-19/Portugal/PT0292/2020\|EPI_ISL_454008\|2020-0 | ···················· | ···················· | ···················· | ···················· | ···················· | ···················· |
| C | 1880 hCoV-19/Portugal/PT0293/2020\|EPI_ISL_454009\|2020-0 | ···················· | ···················· | ···················· | ···················· | ···················· | ···················· |
| C | 1881 hCoV-19/Portugal/PT0294/2020\|EPI_ISL_454010\|2020-0 | ···················· | ···················· | ···················· | ···················· | ···················· | ···················· |
| C | 1882 hCoV-19/Portugal/PT0295/2020\|EPI_ISL_454011\|2020-0 | ···················· | ···················· | ···················· | ···················· | ···················· | ···················· |
| C | 1883 hCoV-19/Portugal/PT0296/2020\|EPI_ISL_454012\|2020-0 | ···················· | ···················· | ···················· | ···················· | ···················· | ···················· |
| C | 1884 hCoV-19/Portugal/PT0297/2020\|EPI_ISL_454013\|2020-0 | ···················· | ···················· | ···················· | ···················· | ···················· | ···················· |
| C | 1885 hCoV-19/Portugal/PT0298/2020\|EPI_ISL_454014\|2020-0 | ···················· | ···················· | ···················· | ···················· | ···················· | ···················· |
| C | 1886 hCoV-19/Portugal/PT0299/2020\|EPI_ISL_454015\|2020-0 | ···················· | ···················· | ···················· | ···················· | ···················· | ···················· |
| C | 1887 hCoV-19/Netherlands/ZuidHolland_114/2020\|EPI_ISL_4 | ···················· | ···················· | ···················· | ···················· | ···················· | ···················· |
| C | 1888 hCoV-19/Netherlands/ZuidHolland_115/2020\|EPI_ISL_4 | ···················· | ···················· | ···················· | ···················· | ···················· | ···················· |
| C | 1889 hCoV-19/Netherlands/ZuidHolland_116/2020\|EPI_ISL_4 | ···················· | ···················· | ···················· | ···················· | ···················· | ···················· |
| C | 1890 hCoV-19/Netherlands/ZuidHolland_117/2020\|EPI_ISL_4 | ···················· | ···················· | ···················· | ···················· | ···················· | ···················· |
| C | 1891 hCoV-19/Netherlands/ZuidHolland_118/2020\|EPI_ISL_4 | ···················· | ···················· | ···················· | ···················· | ···················· | ···················· |
| C | 1893 hCoV-19/India/OR-ILC-RMRC123/2020\|EPI_ISL_455311\|2 | ···················· | ···················· | ···················· | ···················· | ···················· | ···················· |
| C | 1894 hCoV-19/Malaysia/IIUM316/2020\|EPI_ISL_455312\|2020- | ···················· | ···················· | ···················· | ···················· | ···················· | ···················· |
| C | 1895 hCoV-19/Malaysia/IIUM91/2020\|EPI_ISL_455313\|2020-0 | ···················· | ···················· | ···················· | ···················· | ···················· | ···A················ |
| C | 1896 hCoV-19/Spain/AN-ISCIII-201257/2020\|EPI_ISL_455314 | ···················· | ···················· | ···················· | ···················· | ···················· | ···················· |
| C | 1897 hCoV-19/Spain/AN-ISCIII-201488/2020\|EPI_ISL_455315 | ···················· | ···················· | ···················· | ···················· | ···················· | ···················· |
| C | 1898 hCoV-19/Spain/AN-ISCIII-201613/2020\|EPI_ISL_455316 | ···················· | ···················· | ···················· | ···················· | ···················· | ···················· |
| C | 1899 hCoV-19/Spain/AN-ISCIII-201614/2020\|EPI_ISL_455317 | ···················· | ···················· | ···················· | ···················· | ···················· | ···················· |
| C | 1900 hCoV-19/Spain/AN-ISCIII-201615/2020\|EPI_ISL_455318 | ···················· | ···················· | ···················· | ···················· | ···················· | ···················· |
| C | 1901 hCoV-19/Spain/AN-ISCIII-201616/2020\|EPI_ISL_455319 | ···················· | ···················· | ···················· | ···················· | ···················· | ···················· |
| C | 1902 hCoV-19/Spain/AN-ISCIII-201620/2020\|EPI_ISL_455320 | ···················· | ···················· | ···················· | ···················· | ···················· | ···················· |
| C | 1903 hCoV-19/Spain/AN-ISCIII-201622/2020\|EPI_ISL_455321 | ···················· | ···················· | ···················· | ···················· | ···················· | ···················· |
| C | 1904 hCoV-19/Spain/AN-ISCIII-201623/2020\|EPI_ISL_455322 | ···················· | ···················· | ···················· | ···················· | ···················· | ···················· |
| C | 1905 hCoV-19/Spain/AN-ISCIII-201649/2020\|EPI_ISL_455324 | ···················· | ···················· | ···················· | ···················· | ···················· | ···················· |
| C | 1906 hCoV-19/Spain/CN-ISCIII-201811/2020\|EPI_ISL_455325 | ···················· | ···················· | ···················· | ···················· | ···················· | ···················· |
| C | 1907 hCoV-19/Spain/CN-ISCIII-201939/2020\|EPI_ISL_455326 | ···················· | ···················· | ···················· | ···················· | ···················· | ···················· |
| C | 1908 hCoV-19/Spain/CM-ISCIII-201306/2020\|EPI_ISL_455327 | ···················· | ···················· | ···················· | ···················· | ···················· | ···················· |
| C | 1909 hCoV-19/Spain/GA-ISCIII-201792/2020\|EPI_ISL_455328 | ···················· | ···················· | ···················· | ···················· | ···················· | ···················· |
| C | 1910 hCoV-19/Spain/GA-ISCIII-201793/2020\|EPI_ISL_455329 | ···················· | ···················· | ···················· | ···················· | ···················· | ···················· |
| C | 1911 hCoV-19/Spain/GA-ISCIII-201796/2020\|EPI_ISL_455330 | ···················· | ···················· | ···················· | ···················· | ···················· | ···················· |
| C | 1912 hCoV-19/Spain/GA-ISCIII-201863/2020\|EPI_ISL_455331 | ···················· | ···················· | ···················· | ···················· | ···················· | ···················· |
| C | 1913 hCoV-19/Spain/GA-ISCIII-202076/2020\|EPI_ISL_455332 | ···················· | ···················· | ···················· | ···················· | ···················· | ···················· |
| C | 1914 hCoV-19/Spain/GA-ISCIII-202230/2020\|EPI_ISL_455333 | ···················· | ···················· | ···················· | ···················· | ···················· | ···················· |
| C | 1915 hCoV-19/Spain/GA-ISCIII-202282/2020\|EPI_ISL_455334 | ···················· | ···················· | ···················· | ···················· | ···················· | ···················· |
| C | 1916 hCoV-19/Spain/GA-ISCIII-202297/2020\|EPI_ISL_455335 | ···················· | ···················· | ···················· | ···················· | ···················· | ···················· |
| C | 1917 hCoV-19/Spain/RI-ISCIII-201277/2020\|EPI_ISL_455336 | ···················· | ···················· | ···················· | ···················· | ···················· | ···················· |
| C | 1918 hCoV-19/Spain/RI-ISCIII-201334/2020\|EPI_ISL_455337 | ···················· | ···················· | ···················· | ···················· | ···················· | ···················· |
| C | 1919 hCoV-19/Spain/RI-ISCIII-201563/2020\|EPI_ISL_455338 | ···················· | ···················· | ···················· | ···················· | ···················· | ···················· |

```
C 1920 hCoV-19/Spain/RI-ISCIII-201565/2020|EPI_ISL_455339 ···················· ···················· ···················· ···················· ···················· ····················
C 1921 hCoV-19/Spain/RI-ISCIII-201568/2020|EPI_ISL_455340 ···················· ···················· ···················· ···················· ···················· ····················
C 1922 hCoV-19/Spain/RI-ISCIII-201570/2020|EPI_ISL_455341 ···················· ···················· ···················· ···················· ···················· ····················
C 1923 hCoV-19/Spain/RI-ISCIII-201572/2020|EPI_ISL_455342 ···················· ···················· ···················· ···················· ···················· ····················
C 1924 hCoV-19/Spain/RI-ISCIII-201576/2020|EPI_ISL_455343 ···················· ···················· ···················· ···················· ···················· ····················
C 1925 hCoV-19/Spain/ML-ISCIII-202020/2020|EPI_ISL_455344 ···················· ···················· ···················· ···················· ···················· ····················
C 1926 hCoV-19/Spain/ML-ISCIII-202134/2020|EPI_ISL_455345 ···················· ···················· ···················· ···················· ···················· ····················
C 1927 hCoV-19/Spain/ML-ISCIII-202135/2020|EPI_ISL_455346 ···················· ···················· ···················· ···················· ···················· ····················
C 1941 hCoV-19/England/BIRM-5EEAF/2020|EPI_ISL_456661|202 ···················· ···················· ···················· ···················· ···················· ····················
C 1945 hCoV-19/England/BIRM-5F2EE/2020|EPI_ISL_456669|202 ···················· ···················· ···················· ···················· ···················· ····················
C 1947 hCoV-19/England/BIRM-5F363/2020|EPI_ISL_456672|202 ···················· ···················· ···················· ···················· ···················· ····················
C 1949 hCoV-19/England/BIRM-5F3EB/2020|EPI_ISL_456675|202 ···················· ···················· ···················· ···················· ···················· ····················
C 1950 hCoV-19/England/BIRM-5F442/2020|EPI_ISL_456676|202 ···················· ···················· ···················· ···················· ···················· ····················
C 1951 hCoV-19/Kenya/C589/2020|EPI_ISL_457913|2020-04-11 ···················· ···················· ···················· ···················· ···················· ····················
C 1952 hCoV-19/Kenya/C787/2020|EPI_ISL_457921|2020-04-15 ···················· ···················· ···················· ···················· ···················· ····················
C 1953 hCoV-19/Kenya/C926/2020|EPI_ISL_457928|2020-04-17 ···················· ···················· ···················· ···················· ···················· ····················
C 1954 hCoV-19/Oman/RESP-20-1857/2020|EPI_ISL_457937|2020 ···················· ···················· ···················· ···················· ···················· ····················
C 1955 hCoV-19/Oman/RESP-20-1976/2020|EPI_ISL_457938|2020 ···················· ···················· ···················· ···················· ···················· ····················
C 1956 hCoV-19/Oman/RESP-20-2181/2020|EPI_ISL_457939|2020 ···················· ···················· ···················· ···················· ···················· ····················
C 1957 hCoV-19/Uruguay/UY-NYUMC852/2020|EPI_ISL_457948|20 ···················· ···················· ···················· ···················· ···················· ····················
C 1958 hCoV-19/Uruguay/UY-NYUMC856/2020|EPI_ISL_457952|20 ···················· ···················· ···················· ···················· ···················· ····················
C 1959 hCoV-19/Uruguay/UY-NYUMC857/2020|EPI_ISL_457953|20 ···················· ···················· ···················· ···················· ···················· ····················
C 1960 hCoV-19/Uruguay/UY-NYUMC869/2020|EPI_ISL_457965|20 ···················· ···················· ···················· ···················· ···················· ····················
C 1961 hCoV-19/Uruguay/UY-NYUMC876/2020|EPI_ISL_457972|20 ···················· ···················· ···················· ···················· ···················· ····················
C 1962 hCoV-19/Oman/RESP-20-2282/2020|EPI_ISL_457974|2020 ···················· ···················· ···················· ···················· ···················· ····················
C 1963 hCoV-19/Oman/RESP-20-2523/2020|EPI_ISL_457975|2020 ···················· ···················· ···················· ···················· ···················· ····················
C 1964 hCoV-19/Oman/RESP-20-2631/2020|EPI_ISL_457976|2020 ···················· ···················· ···················· ···················· ···················· ····················
C 1965 hCoV-19/Oman/RESP-20-2850/2020|EPI_ISL_457977|2020 ···················· ···················· ···················· ···················· ···················· ····················
C 1966 hCoV-19/Oman/RESP-20-2857/2020|EPI_ISL_457978|2020 ···················· ···················· ···················· ···················· ···················· ····················
C 1967 hCoV-19/Oman/RESP-20-3080/2020|EPI_ISL_457979|2020 ···················· ···················· ···················· ···················· ···················· ····················
C 1968 hCoV-19/Oman/RESP-20-3515/2020|EPI_ISL_457980|2020 ···················· ···················· ···················· ···················· ···················· ····················
C 1969 hCoV-19/Oman/205026358/2020|EPI_ISL_457981|2020-04 ···················· ···················· ···················· ···················· ···················· ····················
C 1970 hCoV-19/Oman/RESP-20-3343/2020|EPI_ISL_457985|2020 ···················· ···················· ···················· ···················· ···················· ····················
C 1971 hCoV-19/Oman/RESP-20-3700/2020|EPI_ISL_457986|2020 ···················· ···················· ···················· ···················· ···················· ····················
C 1972 hCoV-19/Oman/RESP-20-4028/2020|EPI_ISL_457987|2020 ···················· ···················· ···················· ···················· ···················· ····················
C 1973 hCoV-19/Oman/RESP-20-4153/2020|EPI_ISL_457988|2020 ···················· ···················· ···················· ···················· ···················· ····················
C 1974 hCoV-19/Oman/RESP-20-4252/2020|EPI_ISL_457989|2020 ···················· ···················· ···················· ···················· ···················· ····················
C 1975 hCoV-19/Oman/RESP-20-4400/2020|EPI_ISL_457990|2020 ···················· ···················· ···················· ···················· ···················· ····················
C 1976 hCoV-19/Oman/RESP-20-4736/2020|EPI_ISL_457991|2020 ···················· ···················· ···················· ···················· ···················· ····················
C 1977 hCoV-19/Oman/RESP-20-4810/2020|EPI_ISL_457992|2020 ···················· ···················· ···················· ···················· ···················· ····················
C 1978 hCoV-19/Oman/RESP-20-5414/2020|EPI_ISL_457993|2020 ···················· ···················· ···················· ···················· ···················· ····················
C 1979 hCoV-19/Oman/RESP-20-5505/2020|EPI_ISL_457994|2020 ···················· ···················· ···················· ···················· ···················· ····················
C 1980 hCoV-19/Oman/RESP-20-6081/2020|EPI_ISL_457995|2020 ···················· ···················· ···················· ···················· ···················· ····················
C 1981 hCoV-19/Oman/RESP-20-6417/2020|EPI_ISL_457996|2020 ···················· ···················· ···················· ···················· ···················· ····················
C 1982 hCoV-19/Oman/RESP-20-6560/2020|EPI_ISL_457997|2020 ···················· ···················· ···················· ···················· ···················· ····················
C 1983 hCoV-19/Oman/RESP-20-6701/2020|EPI_ISL_457998|2020 ···················· ···················· ···················· ···················· ···················· ····················
C 1984 hCoV-19/Netherlands/NA_665/2020|EPI_ISL_461100|202 ···················· ···················· ···················· ···················· ···················· ····················
C 1985 hCoV-19/Netherlands/NA_666/2020|EPI_ISL_461101|202 ···················· ···················· ···················· ···················· ···················· ····················
C 1986 hCoV-19/Netherlands/NA_667/2020|EPI_ISL_461102|202 ···················· ···················· ···················· ···················· ···················· ····················
C 1987 hCoV-19/Netherlands/NA_668/2020|EPI_ISL_461103|202 ···················· ···················· ···················· ···················· ···················· ····················
C 1988 hCoV-19/Netherlands/NA_669/2020|EPI_ISL_461104|202 ···················· ···················· ···················· ···················· ···················· ····················
C 1989 hCoV-19/Netherlands/NA_670/2020|EPI_ISL_461105|202 ···················· ···················· ···················· ···················· ···················· ····················
C 1990 hCoV-19/Netherlands/NA_671/2020|EPI_ISL_461106|202 ···················· ···················· ···················· ···················· ···················· ····················
C 1991 hCoV-19/Netherlands/NA_673/2020|EPI_ISL_461108|202 ···················· ···················· ···················· ···················· ···················· ····················
C 1992 hCoV-19/Netherlands/NA_674/2020|EPI_ISL_461109|202 ···················· ···················· ···················· ···················· ···················· ····················
C 1993 hCoV-19/Netherlands/NA_676/2020|EPI_ISL_461111|202 ···················· ···················· ···················· ···················· ···················· ····················
C 1994 hCoV-19/Netherlands/NA_677/2020|EPI_ISL_461112|202 ···················· ···················· ···················· ···················· ···················· ····················
C 1995 hCoV-19/Netherlands/NA_678/2020|EPI_ISL_461113|202 ···················· ···················· ···················· ···················· ···················· ····················
C 1996 hCoV-19/Netherlands/NA_679/2020|EPI_ISL_461114|202 ···················· ···················· ···················· ···················· ···················· ····················
C 1997 hCoV-19/Netherlands/NA_680/2020|EPI_ISL_461115|202 ···················· ···················· ···················· ···················· ···················· ····················
C 1998 hCoV-19/Netherlands/NA_681/2020|EPI_ISL_461116|202 ···················· ···················· ···················· ···················· ···················· ····················
C 1999 hCoV-19/Netherlands/NA_683/2020|EPI_ISL_461118|202 ···················· ···················· ···················· ···················· ···················· ····················
C 2000 hCoV-19/Netherlands/NA_685/2020|EPI_ISL_461120|202 ···················· ···················· ···················· ···················· ···················· ····················
C 2001 hCoV-19/Netherlands/NA_686/2020|EPI_ISL_461121|202 ···················· ···················· ···················· ···················· ···················· ····················
C 2002 hCoV-19/Netherlands/NA_687/2020|EPI_ISL_461122|202 ···················· ···················· ···················· ···················· ···················· ····················
C 2003 hCoV-19/Netherlands/NA_688/2020|EPI_ISL_461123|202 ···················· ···················· ···················· ···················· ···················· ····················
C 2004 hCoV-19/Netherlands/NA_690/2020|EPI_ISL_461125|202 ···················· ···················· ···················· ···················· ···················· ····················
C 2005 hCoV-19/Netherlands/NA_691/2020|EPI_ISL_461126|202 ···················· ···················· ···················· ···················· ···················· ····················
C 2006 hCoV-19/Netherlands/NA_692/2020|EPI_ISL_461127|202 ···················· ···················· ···················· ···················· ···················· ····················
C 2007 hCoV-19/Netherlands/NA_693/2020|EPI_ISL_461128|202 ···················· ···················· ···················· ···················· ···················· ····················
C 2008 hCoV-19/Netherlands/NA_694/2020|EPI_ISL_461129|202 ···················· ···················· ···················· ···················· ···················· ····················
C 2009 hCoV-19/Netherlands/NA_695/2020|EPI_ISL_461130|202 ···················· ···················· ···················· ···················· ···················· ····················
C 2010 hCoV-19/Netherlands/NA_696/2020|EPI_ISL_461131|202 ···················· ·····A·············· ···················· ···················· ···················· ····················
C 2011 hCoV-19/Netherlands/NA_697/2020|EPI_ISL_461132|202 ···················· ···················· ···················· ···················· ···················· ····················
C 2012 hCoV-19/Netherlands/NA_698/2020|EPI_ISL_461133|202 ···················· ···················· ···················· ···················· ···················· ····················
C 2013 hCoV-19/Netherlands/NA_699/2020|EPI_ISL_461134|202 ···················· ···················· ···················· ···················· ···················· ····················
C 2014 hCoV-19/Netherlands/NA_700/2020|EPI_ISL_461135|202 ···················· ···················· ···················· ···················· ···················· ····················
C 2015 hCoV-19/Netherlands/NA_701/2020|EPI_ISL_461136|202 ···················· ···················· ···················· ···················· ···················· ····················
```

C 2016 hCoV-19/Netherlands/NA_702/2020|EPI_ISL_461137|202 .................... .................... .................... .................... .................... ....................
C 2017 hCoV-19/Netherlands/NA_704/2020|EPI_ISL_461139|202 .................... .................... .................... .................... .................... ....................
C 2018 hCoV-19/Netherlands/NA_705/2020|EPI_ISL_461140|202 .................... ...A................ .................... .................... .................... ....................
C 2019 hCoV-19/Netherlands/NA_706/2020|EPI_ISL_461141|202 .................... .................... .................... .................... .................... ....................
C 2020 hCoV-19/Netherlands/NA_708/2020|EPI_ISL_461143|202 .................... .................... .................... .................... .................... ....................
C 2021 hCoV-19/Netherlands/NoordBrabant_147/2020|EPI_ISL_ .................... .................... .................... .................... .................... ....................
C 2022 hCoV-19/Netherlands/NoordBrabant_148/2020|EPI_ISL_ .................... .................... .................... .................... .................... ....................
C 2023 hCoV-19/Netherlands/NoordBrabant_149/2020|EPI_ISL_ .................... .................... .................... .................... .................... ....................
C 2024 hCoV-19/Netherlands/NoordBrabant_150/2020|EPI_ISL_ .................... .................... .................... .................... .................... ....................
C 2025 hCoV-19/Netherlands/NoordBrabant_151/2020|EPI_ISL_ .................... .................... .................... .................... .................... ....................
C 2026 hCoV-19/Netherlands/NoordBrabant_153/2020|EPI_ISL_ .................... .................... .................... .................... .................... ....................
C 2027 hCoV-19/Netherlands/NoordBrabant_154/2020|EPI_ISL_ .................... .................... .................... .................... .................... ....................
C 2028 hCoV-19/Netherlands/NoordBrabant_156/2020|EPI_ISL_ .................... .................... .................... .................... .................... ....................
C 2029 hCoV-19/Netherlands/NoordBrabant_157/2020|EPI_ISL_ .................... .................... .................... .................... .................... ....................
C 2030 hCoV-19/Netherlands/NoordBrabant_158/2020|EPI_ISL_ .................... .................... .................... .................... .................... ....................
C 2031 hCoV-19/Netherlands/NoordBrabant_159/2020|EPI_ISL_ .................... .................... .................... .................... .................... ....................
C 2032 hCoV-19/Netherlands/NoordBrabant_160/2020|EPI_ISL_ .................... .................... .................... .................... .................... ....................
C 2033 hCoV-19/Netherlands/NoordBrabant_163/2020|EPI_ISL_ .................... .................... .................... .................... .................... ....................
C 2034 hCoV-19/Netherlands/NoordBrabant_164/2020|EPI_ISL_ .................... .................... .................... .................... .................... ....................
C 2035 hCoV-19/Netherlands/NoordBrabant_165/2020|EPI_ISL_ .................... .................... .................... .................... .................... ....................
C 2036 hCoV-19/Serbia/NS838-04/2020|EPI_ISL_462434|2020-0 .................... .................... .................... .................... .................... ....................
C 2037 hCoV-19/Serbia/KV22-05/2020|EPI_ISL_462435|2020-05 .................... .................... .................... .................... .................... ....................
C 2038 hCoV-19/Serbia/UE38-04/2020|EPI_ISL_462436|2020-04 .................... .................... .................... .................... .................... ....................
C 2039 hCoV-19/Serbia/KV27-04/2020|EPI_ISL_462437|2020-04 .................... .................... .................... .................... .................... ....................
C 2040 hCoV-19/Serbia/KV77-04/2020|EPI_ISL_462438|2020-04 .................... .................... .................... .................... .................... ....................
C 2041 hCoV-19/Japan/TKY9588/2020|EPI_ISL_462439|2020-04 .................... .................... .................... .................... .................... ....................
C 2042 hCoV-19/Japan/TKY9810/2020|EPI_ISL_462440|2020-04 .................... .................... .................... .................... .................... ....................
C 2043 hCoV-19/Japan/TKY10256/2020|EPI_ISL_462441|2020-04 .................... .................... .................... .................... .................... ....................
C 2044 hCoV-19/Japan/TKY10459/2020|EPI_ISL_462442|2020-04 .................... .................... .................... .................... .................... ....................
C 2045 hCoV-19/Japan/TKY10466/2020|EPI_ISL_462443|2020-04 .................... .................... .................... .................... .................... ....................
C 2046 hCoV-19/Japan/TKY10466-2/2020|EPI_ISL_462444|2020- .................... .................... .................... .................... .................... ....................
C 2047 hCoV-19/Japan/TKY10468/2020|EPI_ISL_462445|2020-04 .................... .................... .................... .................... .................... ....................
C 2048 hCoV-19/Japan/TKY3583/2020|EPI_ISL_462446|2020-03 .................... .................... .................... .................... .................... ....................
C 2049 hCoV-19/England/CAMB-1B003B/2020|EPI_ISL_448100|20 .................... .................... .................... .................... .................... ....................
C 2050 hCoV-19/England/CAMB-1B004A/2020|EPI_ISL_448101|20 .................... .................... .................... .................... .................... ....................
C 2051 hCoV-19/England/CAMB-1B0059/2020|EPI_ISL_448102|20 .................... .A.................. .................... .................... .................... ....................
C 2052 hCoV-19/England/CAMB-1B0068/2020|EPI_ISL_448103|20 .................... .................... .................... .................... .................... ...........A........
C 2053 hCoV-19/England/CAMB-1B0086/2020|EPI_ISL_448105|20 .................... .................... .................... .................... .................... ....................
C 2054 hCoV-19/England/CAMB-1B0095/2020|EPI_ISL_448106|20 .................... .................... .................... .................... .................... ....................
C 2055 hCoV-19/England/CAMB-1B00A4/2020|EPI_ISL_448107|20 .................... .................... .................... .................... .................... ....................
C 2056 hCoV-19/England/CAMB-1B00D1/2020|EPI_ISL_448108|20 .................... .................... .................... .................... .................... ....................
C 2057 hCoV-19/England/CAMB-1B00FF/2020|EPI_ISL_448109|20 .................... .................... .................... .................... .................... ....................
C 2058 hCoV-19/England/CAMB-1B011A/2020|EPI_ISL_448111|20 .................... .................... .................... .................... .................... ....................
C 2059 hCoV-19/England/CAMB-1B05D2/2020|EPI_ISL_448114|20 .................... .................... .................... .................... .................... ....................
C 2060 hCoV-19/Scotland/CVR1501/2020|EPI_ISL_448117|2020- .................... .................... .................... .................... .................... ....................
C 2061 hCoV-19/Scotland/CVR1584/2020|EPI_ISL_448118|2020- .................... .A.................. .................... .................... .................... ....................
C 2062 hCoV-19/Scotland/CVR1601/2020|EPI_ISL_448119|2020- .................... ...................A .................... .................... .................... .................... 1
C 2063 hCoV-19/Scotland/CVR1623/2020|EPI_ISL_448120|2020- .................... .................... .................... .................... .................... ....................
C 2064 hCoV-19/Scotland/CVR1624/2020|EPI_ISL_448121|2020- .................... .A.................. .................... .................... .................... ....................
C 2065 hCoV-19/Scotland/CVR1641/2020|EPI_ISL_448122|2020- .................... .................... .................... .................... .................... ....................
C 2066 hCoV-19/Scotland/CVR1642/2020|EPI_ISL_448123|2020- .................... .................... .................... .................... .................... ....................
C 2067 hCoV-19/Scotland/CVR1679/2020|EPI_ISL_448124|2020- .................... .................... .................... .................... .................... ....................
C 2068 hCoV-19/Scotland/CVR1701/2020|EPI_ISL_448126|2020- .................... .................... .................... .................... .................... ....................
C 2069 hCoV-19/Scotland/CVR1713/2020|EPI_ISL_448127|2020- .................... ...................A .................... .................... .................... .................... 1
C 2070 hCoV-19/Scotland/CVR1718/2020|EPI_ISL_448128|2020- .................... ...................A .................... .................... .................... .................... 1
C 2071 hCoV-19/Scotland/CVR1805/2020|EPI_ISL_448129|2020- .................... .................... .................... .................... .................... ....................
C 2072 hCoV-19/Scotland/CVR1806/2020|EPI_ISL_448130|2020- .................... .................... .................... .................... .................... ....................
C 2073 hCoV-19/Scotland/CVR1822/2020|EPI_ISL_448131|2020- .................... ...................A .................... .................... .................... .................... 1
C 2074 hCoV-19/Scotland/CVR1825/2020|EPI_ISL_448132|2020- .................... .................... .................... .................... .................... ....................
C 2075 hCoV-19/Scotland/CVR1826/2020|EPI_ISL_448133|2020- ..................T. ...................A .................... .................... .................... .................... 1
C 2076 hCoV-19/Scotland/CVR1840/2020|EPI_ISL_448134|2020- .................... .................... .................... .................... .................... ....................
C 2077 hCoV-19/Scotland/CVR1902/2020|EPI_ISL_448136|2020- .................... .................... .................... .................... .................... ....................
C 2078 hCoV-19/Scotland/CVR1927/2020|EPI_ISL_448138|2020- .................... .................... .................... .................... .................... ....................
C 2079 hCoV-19/Scotland/CVR1953/2020|EPI_ISL_448139|2020- .................... .A.................. .................... .................... .................... ....................
C 2080 hCoV-19/Scotland/CVR2329/2020|EPI_ISL_448180|2020- .................... .................... .................... .................... .................... ....................
C 2081 hCoV-19/Scotland/CVR2345/2020|EPI_ISL_448182|2020- .................... .................... .................... .................... .................... ....................
C 2082 hCoV-19/Scotland/CVR2388/2020|EPI_ISL_448183|2020- .................... .................... .................... .................... .................... ....................
C 2083 hCoV-19/Scotland/CVR2414/2020|EPI_ISL_448184|2020- .................... .................... .................... .................... .................... ....................
C 2084 hCoV-19/Scotland/CVR2415/2020|EPI_ISL_448185|2020- .................... .A.................. .................... .................... .................... ....................
C 2085 hCoV-19/Scotland/CVR2433/2020|EPI_ISL_448187|2020- .................... .A.................. .................... .................... .................... ....................
C 2086 hCoV-19/Scotland/CVR2526/2020|EPI_ISL_448188|2020- .................... .A.................. .................... .................... .................... ....................
C 2087 hCoV-19/Scotland/CVR2686/2020|EPI_ISL_448189|2020- .................... ..........A......... .................... .................... .................... ....................
C 2088 hCoV-19/Scotland/CVR2742/2020|EPI_ISL_448190|2020- .................... .A.................. .................... .................... .................... ....................
C 2089 hCoV-19/Scotland/CVR2753/2020|EPI_ISL_448191|2020- .................... .................... .................... .................... .................... ....................
C 2090 hCoV-19/Scotland/CVR2768/2020|EPI_ISL_448192|2020- .................... .................... .................... .................... .................... ....................
C 2091 hCoV-19/Scotland/CVR2771/2020|EPI_ISL_448193|2020- .................... .................... .................... .................... .................... ....................
C 2092 hCoV-19/Scotland/CVR2817/2020|EPI_ISL_448195|2020- .................... ...................A .................... .................... .................... .................... 1
C 2093 hCoV-19/Scotland/CVR2840/2020|EPI_ISL_448196|2020- .................... .................... .................... .................... .................... ....................

C 2094 hCoV-19/Scotland/CVR2842/2020|EPI_ISL_448197|2020- ···················· ···················A ···················· ···················· ···················· ···················· 1
C 2095 hCoV-19/Scotland/CVR2852/2020|EPI_ISL_448198|2020- ···················· ···················· ···················· ···················· ···················· ····················
C 2096 hCoV-19/Scotland/CVR2853/2020|EPI_ISL_448199|2020- ···················· ···················· ···················· ···················· ···················· ····················
C 2097 hCoV-19/USA/NY-NYUMC683/2020|EPI_ISL_450400|2020-0 ···················· ···················· ···················· ···················· ···················· ····················
C 2098 hCoV-19/USA/NY-NYUMC684/2020|EPI_ISL_450401|2020-0 ···················· ···················· ···················· ···················· ···················· ····················
C 2099 hCoV-19/USA/NY-NYUMC685/2020|EPI_ISL_450402|2020-0 ···················· ···················· ···················· ···················· ···················· ····················
C 2100 hCoV-19/USA/CA-CZB-1196/2020|EPI_ISL_450445|2020-0 ···················· ···················· ···················· ···················· ···················· ····················
C 2101 hCoV-19/USA/CA-CZB-1197/2020|EPI_ISL_450446|2020-0 ···················· ···················· ···················· ···················· ···················· ····················
C 2102 hCoV-19/USA/CA-CZB-1198/2020|EPI_ISL_450447|2020-0 ···················· ···················· ···················· ···················· ···················· ····················
C 2103 hCoV-19/USA/CA-CZB-1199/2020|EPI_ISL_450448|2020-0 ···················· ···················· ···················· ···················· ···················· ····················
C 2104 hCoV-19/USA/CA-CZB-1200/2020|EPI_ISL_450449|2020-0 ···················· ···················· ···················· ···················· ···················· ····················
C 2105 hCoV-19/USA/CA-CZB-1201/2020|EPI_ISL_450450|2020-0 ···················· ···················· ···················· ···················· ···················· ····················
C 2106 hCoV-19/USA/CA-CZB-1202/2020|EPI_ISL_450451|2020-0 ···················· ···················· ···················· ···················· ···················· ····················
C 2107 hCoV-19/Lithuania/ChVir2225/2020|EPI_ISL_450496|20 ···················· ···················· ···················· ···················· ···················· ····················
C 2108 hCoV-19/Lithuania/ChVir2226/2020|EPI_ISL_450497|20 ···················· ···················· ···················· ···················· ···················· ····················
C 2109 hCoV-19/Estonia/ChVir1984/2020|EPI_ISL_450498|2020 ···················· ···················· ···················· ···················· ···················· ····················
C 2110 hCoV-19/Switzerland/BE-ETHZ-100117/2020|EPI_ISL_45 ···················· ···················· ···················· ···················· ···················· ····················
C 2111 hCoV-19/Switzerland/BE-ETHZ-100118/2020|EPI_ISL_45 ···················· ···················· ···················· ···················· ···················· ····················
C 2115 hCoV-19/Switzerland/BE-ETHZ-100122/2020|EPI_ISL_45 ···················· ···················· ···················· ···················· ···················· ····················
C 2117 hCoV-19/Switzerland/JU-ETHZ-100124/2020|EPI_ISL_45 ···················· ···················· ···················· ···················· ·······S············ ····················
C 2118 hCoV-19/Switzerland/AG-ETHZ-100125/2020|EPI_ISL_45 ···················· ···················· ···················· ···················· ···················· ····················
C 2119 hCoV-19/Switzerland/TI-ETHZ-100127/2020|EPI_ISL_45 ···················· ···················· ···················· ···················· ···················· ····················
C 2120 hCoV-19/Switzerland/VD-ETHZ-100128/2020|EPI_ISL_45 ···················· ···················A ···················· ···················· ···················· ·R·················· 1
C 2121 hCoV-19/Switzerland/BS-ETHZ-100129/2020|EPI_ISL_45 ···················· ···················· ···················· ···················· ···················· ····················
C 2123 hCoV-19/Switzerland/BS-ETHZ-100132/2020|EPI_ISL_45 ···················· ···················· ···················· ···················· ···················· ····················
C 2124 hCoV-19/Switzerland/AG-ETHZ-100133/2020|EPI_ISL_45 ···················· ···················· ···················· ···················· ···················· ····················
C 2129 hCoV-19/Switzerland/GR-ETHZ-100138/2020|EPI_ISL_45 ···················· ···················· ···················· ···················· ···················· ····················
C 2130 hCoV-19/Switzerland/GR-ETHZ-100139/2020|EPI_ISL_45 ···················· ···················· ···················· ···················· ···················· ····················
C 2132 hCoV-19/Switzerland/GR-ETHZ-100141/2020|EPI_ISL_45 ···················· ···················· ···················· ···················· ·······S············ ····················
C 2134 hCoV-19/Switzerland/BS-ETHZ-100144/2020|EPI_ISL_45 ···················· ···················· ···················· ···················· ···················· ····················
C 2135 hCoV-19/Switzerland/TI-ETHZ-100146/2020|EPI_ISL_45 ···················· ···················· ···················· ···················· ···················· ····················
C 2137 hCoV-19/Switzerland/BS-ETHZ-100148/2020|EPI_ISL_45 ···················· ···················· ···················· ···················· ···················· ····················
C 2138 hCoV-19/Switzerland/GR-ETHZ-100150/2020|EPI_ISL_45 ···················· ···················· ···················· ···················· ···················· ····················
C 2139 hCoV-19/Switzerland/SG-ETHZ-100152/2020|EPI_ISL_45 ···················· ···················· ···················· ···················· ···················· ····················
C 2140 hCoV-19/Switzerland/BE-ETHZ-100154/2020|EPI_ISL_45 ···················· ···················· ···················· ···················· ···················· ····················
C 2141 hCoV-19/Switzerland/GR-ETHZ-100790/2020|EPI_ISL_45 ···················· ···················· ···················· ···················· ···················· ····················
C 2144 hCoV-19/Switzerland/VD-ETHZ-100794/2020|EPI_ISL_45 ···················· ···················· ···················· ···················· ···················· ····················
C 2145 hCoV-19/Switzerland/VD-ETHZ-100795/2020|EPI_ISL_45 ···················· ···················· ···················· ···················· ···················· ····················
C 2147 hCoV-19/Switzerland/ZH-ETHZ-100797/2020|EPI_ISL_45 ···················· ···················· ···················· ···················· ···················· ····················
C 2148 hCoV-19/Switzerland/VS-ETHZ-100798/2020|EPI_ISL_45 ···················· ·A·················· ···················· ···················· ···········NN······· ····················
C 2149 hCoV-19/Switzerland/UR-ETHZ-100799/2020|EPI_ISL_45 ···················· ···················· ···················· ···················· ···················· ····················
C 2150 hCoV-19/Switzerland/BL-ETHZ-100800/2020|EPI_ISL_45 ···················· ···················· ···················· ···················· ···················· ····················
C 2151 hCoV-19/Switzerland/BE-ETHZ-100801/2020|EPI_ISL_45 ···················· ···················· ···················· ···················· ···················· ····················
C 2155 hCoV-19/Switzerland/VD-ETHZ-100805/2020|EPI_ISL_45 ···················· ···················· ···················· ···················· ···················· ····················
C 2158 hCoV-19/Switzerland/AG-ETHZ-100809/2020|EPI_ISL_45 ···················· ···················· ···················· ···················· ···················· ····················
C 2159 hCoV-19/Switzerland/GR-ETHZ-100810/2020|EPI_ISL_45 ···················· ···················· ···················· ···················· ···················· ···········Y········
C 2161 hCoV-19/Switzerland/GR-ETHZ-100813/2020|EPI_ISL_45 ···················· ···················· ···················· ···················· ···················· ····················
C 2162 hCoV-19/Switzerland/BL-ETHZ-100814/2020|EPI_ISL_45 ···················· ···················· ···················· ···················· ···················· ····················
C 2164 hCoV-19/Switzerland/BL-ETHZ-100816/2020|EPI_ISL_45 ···················· ···················· ···················· ···················· ···················· ····················
C 2165 hCoV-19/Switzerland/ZH-ETHZ-100817/2020|EPI_ISL_45 ···················· ···················· ···················· ···················· ·······S············ ····················
C 2166 hCoV-19/Switzerland/VD-ETHZ-100820/2020|EPI_ISL_45 ···················· ···················· ···················· ···················· ·······S············ ····················
C 2167 hCoV-19/Switzerland/VD-ETHZ-100821/2020|EPI_ISL_45 ···················· ···················· ···················· ···················· ···················· ····················
C 2171 hCoV-19/Switzerland/BL-ETHZ-100827/2020|EPI_ISL_45 ···················· ···················· ···················· ···················· ···················· ····················
C 2174 hCoV-19/Switzerland/UR-ETHZ-100832/2020|EPI_ISL_45 ···················· ···················· ···················· ···················· ·······S············ ····················
C 2175 hCoV-19/Switzerland/BL-ETHZ-100833/2020|EPI_ISL_45 ···················· ···················· ···················· ···················· ·······S············ ····················
C 2177 hCoV-19/India/MH-NIV-9127/2020|EPI_ISL_454556|2020 ···················· ···················· ···················· ···················· ···················· ····················
C 2178 hCoV-19/India/MH-NIV-QC-795/2020|EPI_ISL_454566|20 ···················· ···················· ···················· ···················· ···················· ····················
C 2180 hCoV-19/India/MH-NIV-QC-798/2020|EPI_ISL_454568|20 ···················· ···················· ···················· ···················· ···················· ····················
C 2183 hCoV-19/Kazakhstan/7263/2020|EPI_ISL_454571|2020-0 ···················· ···················· ···················· ···················· ···················· ····················
C 2184 hCoV-19/Kazakhstan/7341/2020|EPI_ISL_454572|2020-0 ···················· ···················· ···················· ···················· ···················· ····················
C 2185 hCoV-19/Croatia/OY-S1new/2020|EPI_ISL_454574|2020- ···················· ···················· ···················· ···················· ···················· ····················
C 2186 hCoV-19/Kazakhstan/38716/2020|EPI_ISL_454575|2020- ···················· ···················· ···················· ···················· ···················· ····················
C 2187 hCoV-19/Kazakhstan/16183/2020|EPI_ISL_454576|2020- ···················· ···················· ···················· ···················· ···················· ····················
C 2188 hCoV-19/Netherlands/NA_424/2020|EPI_ISL_455226|202 ···················· ···················· ···················· ···················· ···················· ····················
C 2189 hCoV-19/Netherlands/NA_425/2020|EPI_ISL_455227|202 ···················· ···················· ···················· ···················· ···················· ····················
C 2190 hCoV-19/Netherlands/NA_426/2020|EPI_ISL_455228|202 ···················· ···················· ···················· ···················· ···················· ····················
C 2191 hCoV-19/Netherlands/NA_429/2020|EPI_ISL_455231|202 ···················· ···················· ···················· ···················· ···················· ····················
C 2192 hCoV-19/Netherlands/NA_430/2020|EPI_ISL_455232|202 ···················· ···················· ···················· ···················· ···················· ····················
C 2193 hCoV-19/Netherlands/NA_431/2020|EPI_ISL_455233|202 ···················· ···················· ···················· ···················· ···················· ····················
C 2194 hCoV-19/Netherlands/NA_433/2020|EPI_ISL_455235|202 ···················· ···················· ···················· ···················· ···················· ····················
C 2195 hCoV-19/Netherlands/NA_434/2020|EPI_ISL_455236|202 ···················· ···················· ···················· ···················· ···················· ····················
C 2196 hCoV-19/Netherlands/NA_435/2020|EPI_ISL_455237|202 ···················· ···················· ···················· ···················· ···················· ····················
C 2197 hCoV-19/Netherlands/NA_436/2020|EPI_ISL_455238|202 ···················· ···················· ···················· ···················· ···················· ····················
C 2198 hCoV-19/Netherlands/NA_439/2020|EPI_ISL_455241|202 ···················· ···················· ···················· ···················· ···················· ····················
C 2199 hCoV-19/Netherlands/NA_440/2020|EPI_ISL_455242|202 ···················· ···················· ···················· ···················· ···················· ····················
C 2200 hCoV-19/Netherlands/NA_442/2020|EPI_ISL_455244|202 ···················· ···················· ···················· ···················· ···················· ····················
C 2201 hCoV-19/Netherlands/NA_443/2020|EPI_ISL_455245|202 ···················· ···················· ···················· ···················· ···················· ····················
C 2202 hCoV-19/Netherlands/NA_444/2020|EPI_ISL_455246|202 ···················· ···················· ···················· ···················· ···················· ····················

| | | | | | | | | |
|---|---|---|---|---|---|---|---|---|
| C | 2203 | hCoV-19/Netherlands/NA_447/2020\|EPI_ISL_455249\|202 | .................... | .................... | .................... | .................... | .................... | .................... |
| C | 2204 | hCoV-19/Netherlands/NA_449/2020\|EPI_ISL_455251\|202 | .................... | .................... | .................... | .................... | .................... | .................... |
| C | 2205 | hCoV-19/Netherlands/NA_450/2020\|EPI_ISL_455252\|202 | .................... | .................... | .................... | .................... | .................... | .................... |
| C | 2206 | hCoV-19/Netherlands/NA_451/2020\|EPI_ISL_455253\|202 | .................... | .................... | .................... | .................... | .................... | .................... |
| C | 2207 | hCoV-19/Netherlands/NA_452/2020\|EPI_ISL_455254\|202 | .................... | .................... | .................... | .................... | .................... | .................... |
| C | 2208 | hCoV-19/Netherlands/NA_453/2020\|EPI_ISL_455255\|202 | .................... | .................... | .................... | .................... | .................... | .................... |
| C | 2209 | hCoV-19/Netherlands/NA_454/2020\|EPI_ISL_455256\|202 | .................... | .................... | .................... | .................... | .................... | .................... |
| C | 2210 | hCoV-19/Netherlands/NA_455/2020\|EPI_ISL_455257\|202 | .................... | .................... | .................... | .................... | .................... | .................... |
| C | 2211 | hCoV-19/Netherlands/NA_456/2020\|EPI_ISL_455258\|202 | .................... | .................... | .................... | .................... | .................... | .................... |
| C | 2212 | hCoV-19/Netherlands/NA_457/2020\|EPI_ISL_455259\|202 | .................... | .................... | .................... | .................... | .................... | .................... |
| C | 2213 | hCoV-19/Netherlands/NA_458/2020\|EPI_ISL_455260\|202 | .................... | .................... | .................... | .................... | .................... | .................... |
| C | 2214 | hCoV-19/Netherlands/NA_459/2020\|EPI_ISL_455261\|202 | .................... | .................... | .................... | .................... | .................... | .................... |
| C | 2215 | hCoV-19/Netherlands/NA_460/2020\|EPI_ISL_455262\|202 | .................... | .................... | .................... | .................... | .................... | .................... |
| C | 2216 | hCoV-19/Netherlands/NA_461/2020\|EPI_ISL_455263\|202 | .................... | .................... | .................... | .................... | .................... | .................... |
| C | 2217 | hCoV-19/Netherlands/NA_464/2020\|EPI_ISL_455266\|202 | .................... | .................... | .................... | .................... | .................... | .................... |
| C | 2218 | hCoV-19/Netherlands/NA_467/2020\|EPI_ISL_455269\|202 | .................... | .................... | .................... | .................... | .................... | .................... |
| C | 2219 | hCoV-19/Netherlands/NA_468/2020\|EPI_ISL_455270\|202 | .................... | .................... | .................... | .................... | .................... | .................... |
| C | 2220 | hCoV-19/Netherlands/NA_472/2020\|EPI_ISL_455274\|202 | .................... | .................... | .................... | .................... | .................... | .................... |
| C | 2221 | hCoV-19/Netherlands/NA_473/2020\|EPI_ISL_455275\|202 | .................... | .................... | .................... | .................... | .................... | .................... |
| C | 2222 | hCoV-19/Netherlands/NA_474/2020\|EPI_ISL_455276\|202 | .................... | .................... | .................... | .................... | .................... | .................... |
| C | 2223 | hCoV-19/Netherlands/NA_475/2020\|EPI_ISL_455277\|202 | .................... | .................... | .................... | .................... | .................... | .................... |
| C | 2224 | hCoV-19/Netherlands/NA_476/2020\|EPI_ISL_455278\|202 | .................... | .................... | .................... | .................... | .................... | .................... |
| C | 2225 | hCoV-19/Netherlands/NA_477/2020\|EPI_ISL_455279\|202 | .................... | .................... | .................... | .................... | .................... | .................... |
| C | 2226 | hCoV-19/Netherlands/NA_478/2020\|EPI_ISL_455280\|202 | .................... | .................... | .................... | .................... | .................... | .................... |
| C | 2227 | hCoV-19/Netherlands/NA_479/2020\|EPI_ISL_455281\|202 | .................... | .................... | .................... | .................... | .................... | .................... |
| C | 2228 | hCoV-19/Netherlands/NA_481/2020\|EPI_ISL_455283\|202 | .................... | .................... | .................... | .................... | .................... | .................... |
| C | 2229 | hCoV-19/Netherlands/NA_482/2020\|EPI_ISL_455284\|202 | .................... | .................... | .................... | .................... | .................... | .................... |
| C | 2230 | hCoV-19/Netherlands/NA_483/2020\|EPI_ISL_455285\|202 | .................... | .................... | .................... | .................... | .................... | .................... |
| C | 2231 | hCoV-19/Netherlands/NA_484/2020\|EPI_ISL_455286\|202 | .................... | .................... | .................... | .................... | .................... | .................... |
| C | 2232 | hCoV-19/Netherlands/NA_485/2020\|EPI_ISL_455287\|202 | .................... | .................... | .................... | .................... | .................... | .................... |
| C | 2233 | hCoV-19/Netherlands/NA_592/2020\|EPI_ISL_455289\|202 | .................... | .................... | .................... | .................... | .................... | .................... |
| C | 2234 | hCoV-19/Netherlands/NoordBrabant_106/2020\|EPI_ISL_ | .................... | .................... | .................... | .................... | .................... | .................... |
| C | 2235 | hCoV-19/Netherlands/NoordBrabant_107/2020\|EPI_ISL_ | .................... | .................... | .................... | .................... | .................... | .................... |
| C | 2236 | hCoV-19/Netherlands/Utrecht_24/2020\|EPI_ISL_455292 | .................... | .................... | .................... | .................... | .................... | .................... |
| C | 2237 | hCoV-19/Netherlands/Utrecht_25/2020\|EPI_ISL_455293 | .................... | .................... | .................... | .................... | .................... | .................... |
| C | 2238 | hCoV-19/Netherlands/ZuidHolland_107/2020\|EPI_ISL_4 | .................... | .................... | .................... | .................... | .................... | .................... |
| C | 2239 | hCoV-19/Netherlands/ZuidHolland_108/2020\|EPI_ISL_4 | .................... | .................... | .................... | .................... | .................... | .................... |
| C | 2240 | hCoV-19/Netherlands/ZuidHolland_109/2020\|EPI_ISL_4 | .................... | .................... | .................... | .................... | .................... | .................... |
| C | 2241 | hCoV-19/Netherlands/ZuidHolland_110/2020\|EPI_ISL_4 | .................... | .................... | .................... | .................... | .................... | .................... |
| C | 2242 | hCoV-19/Sweden/20-50249/2020\|EPI_ISL_455888\|2020-0 | .................... | .................... | .................... | .................... | .................... | .................... |
| C | 2243 | hCoV-19/Sweden/20-50250/2020\|EPI_ISL_455889\|2020-0 | .................... | .................... | .................... | .................... | .................... | .................... |
| C | 2244 | hCoV-19/Sweden/20-50259/2020\|EPI_ISL_455897\|2020-0 | .................... | .................... | .................... | .................... | .................... | .................... |
| C | 2245 | hCoV-19/Sweden/20-50284/2020\|EPI_ISL_455898\|2020-0 | .................... | .................... | .................... | .................... | .................... | .................... |
| C | 2246 | hCoV-19/Sweden/20-50286/2020\|EPI_ISL_455899\|2020-0 | .................... | .................... | .................... | .................... | .................... | .................... |
| C | 2253 | hCoV-19/Australia/VIC1664/2020\|EPI_ISL_456565\|2020 | .................... | .................... | .................... | .................... | .................... | .................... |
| C | 2265 | hCoV-19/Kenya/C3129/2020\|EPI_ISL_457897\|2020-04-28 | .................... | .................... | .................... | .................... | .................... | .................... |
| C | 2266 | hCoV-19/Kenya/C3427/2020\|EPI_ISL_457906\|2020-04-29 | .................... | .................... | .................... | .................... | .................... | .................... |
| C | 2267 | hCoV-19/Kenya/C656/2020\|EPI_ISL_457915\|2020-04-13 | .................... | .................... | .................... | .................... | .................... | .................... |
| C | 2268 | hCoV-19/Kenya/C713/2020\|EPI_ISL_457920\|2020-04-14 | .................... | .................... | .................... | .................... | .................... | .................... |
| C | 2269 | hCoV-19/Netherlands/NA_709/2020\|EPI_ISL_461144\|202 | .................... | .................... | .................... | .................... | .................... | .................... |
| C | 2270 | hCoV-19/Netherlands/NA_710/2020\|EPI_ISL_461145\|202 | .................... | .................... | .................... | .................... | .................... | .................... |
| C | 2271 | hCoV-19/Netherlands/NoordBrabant_110/2020\|EPI_ISL_ | .................... | .................... | .................... | .................... | .................... | .................... |
| C | 2272 | hCoV-19/Netherlands/NoordBrabant_111/2020\|EPI_ISL_ | .................... | .................... | .................... | .................... | .................... | .................... |
| C | 2273 | hCoV-19/Netherlands/NoordBrabant_113/2020\|EPI_ISL_ | .................... | .................... | .................... | .................... | .................... | .................... |
| C | 2274 | hCoV-19/Netherlands/NoordBrabant_114/2020\|EPI_ISL_ | .................... | .................... | .................... | .................... | .................... | .................... |
| C | 2275 | hCoV-19/Netherlands/NoordBrabant_115/2020\|EPI_ISL_ | .................... | .................... | .................... | .................... | .................... | .................... |
| C | 2276 | hCoV-19/Netherlands/NoordBrabant_116/2020\|EPI_ISL_ | .................... | .................... | .................... | .................... | .................... | .................... |
| C | 2277 | hCoV-19/Netherlands/NoordBrabant_117/2020\|EPI_ISL_ | .................... | .................... | .................... | .................... | .................... | .................... |
| C | 2278 | hCoV-19/Netherlands/NoordBrabant_118/2020\|EPI_ISL_ | .................... | .................... | .................... | .................... | .................... | .................... |
| C | 2279 | hCoV-19/Netherlands/NoordBrabant_120/2020\|EPI_ISL_ | .................... | .................... | .................... | .................... | .................... | .................... |
| C | 2280 | hCoV-19/Netherlands/NoordBrabant_121/2020\|EPI_ISL_ | .................... | .................... | .................... | .................... | .................... | .................... |
| C | 2281 | hCoV-19/Netherlands/NoordBrabant_122/2020\|EPI_ISL_ | .................... | .................... | .................... | .................... | .................... | .................... |
| C | 2282 | hCoV-19/Netherlands/NoordBrabant_123/2020\|EPI_ISL_ | .................... | .................... | .................... | .................... | .................... | .................... |
| C | 2283 | hCoV-19/Netherlands/NoordBrabant_124/2020\|EPI_ISL_ | .................... | .................... | .................... | .................... | .................... | .................... |
| C | 2284 | hCoV-19/Netherlands/NoordBrabant_125/2020\|EPI_ISL_ | .................... | .................... | .................... | .................... | .................... | .................... |
| C | 2285 | hCoV-19/Netherlands/NoordBrabant_126/2020\|EPI_ISL_ | .................... | .................... | .................... | .................... | .................... | .................... |
| C | 2286 | hCoV-19/Netherlands/NoordBrabant_127/2020\|EPI_ISL_ | .................... | .................... | .................... | .................... | .................... | .................... |
| C | 2287 | hCoV-19/Netherlands/NoordBrabant_128/2020\|EPI_ISL_ | .................... | .................... | .................... | .................... | .................... | .................... |
| C | 2288 | hCoV-19/Netherlands/NoordBrabant_129/2020\|EPI_ISL_ | .................... | .................... | .................... | .................... | .................... | .................... |
| C | 2289 | hCoV-19/Netherlands/NoordBrabant_130/2020\|EPI_ISL_ | .................... | .................... | .................... | .................... | .................... | .................... |
| C | 2290 | hCoV-19/Netherlands/NoordBrabant_132/2020\|EPI_ISL_ | .................... | .................... | .................... | .................... | .................... | .................... |
| C | 2291 | hCoV-19/Netherlands/NoordBrabant_133/2020\|EPI_ISL_ | .................... | .................... | .................... | .................... | .................... | .................... |
| C | 2292 | hCoV-19/Netherlands/NoordBrabant_134/2020\|EPI_ISL_ | .................... | .................... | .................... | .................... | .................... | .................... |
| C | 2293 | hCoV-19/Netherlands/NoordBrabant_136/2020\|EPI_ISL_ | .................... | .................... | .................... | .................... | .................... | .................... |
| C | 2294 | hCoV-19/Netherlands/NoordBrabant_137/2020\|EPI_ISL_ | .................... | .................... | .................... | .................... | .................... | .................... |
| C | 2295 | hCoV-19/Netherlands/NoordBrabant_138/2020\|EPI_ISL_ | .................... | .................... | .................... | .................... | .................... | .................... |
| C | 2296 | hCoV-19/Netherlands/NoordBrabant_139/2020\|EPI_ISL_ | .................... | .................... | .................... | .................... | .................... | .................... |
| C | 2297 | hCoV-19/Netherlands/NoordBrabant_140/2020\|EPI_ISL_ | .................... | .................... | .................... | .................... | .................... | .................... |

| | | | | | | | | |
|---|---|---|---|---|---|---|---|---|
| C | 2298 | hCoV-19/Netherlands/NoordBrabant_141/2020\|EPI_ISL_ | ···················· | ···················· | ···················· | ···················· | ···················· | ···················· |
| C | 2299 | hCoV-19/Netherlands/NoordBrabant_142/2020\|EPI_ISL_ | ···················· | ···················· | ···················· | ···················· | ···················· | ···················· |
| C | 2300 | hCoV-19/Netherlands/NoordBrabant_143/2020\|EPI_ISL_ | ···················· | ···················· | ···················· | ···················· | ···················· | ···················· |
| C | 2301 | hCoV-19/Netherlands/NoordBrabant_144/2020\|EPI_ISL_ | ···················· | ···················· | ···················· | ···················· | ···················· | ···················· |
| C | 2302 | hCoV-19/Netherlands/NoordBrabant_145/2020\|EPI_ISL_ | ·····T·············· | ···················· | ···················· | ···················· | ···················· | ···················· |
| C | 2303 | hCoV-19/Netherlands/NoordBrabant_146/2020\|EPI_ISL_ | ···················· | ···················· | ···················· | ···················· | ···················· | ···················· |
| C | 2304 | hCoV-19/Egypt/CUNCI-HGC5I015/2020\|EPI_ISL_475722\|2 | ···················· | ···················· | ···················· | ···················· | ···················· | ···················· |
| C | 2305 | hCoV-19/Israel/2107681/2020\|EPI_ISL_447350\|2020-04 | ···················· | ···················· | ···················· | ···················· | ···················· | ···················· |
| C | 2306 | hCoV-19/Israel/2107137/2020\|EPI_ISL_447351\|2020-04 | ···················· | ···················· | ···················· | ···················· | ···················· | ···················· |
| C | 2307 | hCoV-19/Israel/2107132/2020\|EPI_ISL_447352\|2020-04 | ···················· | ···················· | ···················· | ···················· | ···················· | ···················· |
| C | 2308 | hCoV-19/Israel/2099421/2020\|EPI_ISL_447353\|2020-04 | ···················· | ···················· | ···················· | ···················· | ···················· | ···················· |
| C | 2309 | hCoV-19/Israel/2099251/2020\|EPI_ISL_447355\|2020-04 | ···················· | ···················· | ···················· | ···················· | ···················· | ···················· |
| C | 2310 | hCoV-19/Israel/2099159/2020\|EPI_ISL_447356\|2020-04 | ···················· | ···················· | ···················· | ···················· | ···················· | ···················· |
| C | 2311 | hCoV-19/Israel/2099019/2020\|EPI_ISL_447357\|2020-04 | ···················· | ···················· | ···················· | ···················· | ···················· | ···················· |
| C | 2312 | hCoV-19/Israel/2089866/2020\|EPI_ISL_447359\|2020-04 | ···················· | ···················· | ···················· | ···················· | ···················· | ···················· |
| C | 2313 | hCoV-19/Israel/2089863/2020\|EPI_ISL_447360\|2020-04 | ···················· | ···················· | ···················· | ···················· | ···················· | ···················· |
| C | 2314 | hCoV-19/Israel/2089861/2020\|EPI_ISL_447361\|2020-04 | ···················· | ···················· | ···················· | ···················· | ···················· | ···················· |
| C | 2315 | hCoV-19/Israel/2089812/2020\|EPI_ISL_447364\|2020-04 | ···················· | ···················· | ···················· | ···················· | ···················· | ···················· |
| C | 2316 | hCoV-19/Israel/2089723/2020\|EPI_ISL_447365\|2020-04 | ···················· | ···················· | ···················· | ···················· | ···················· | ···················· |
| C | 2317 | hCoV-19/Israel/2089718/2020\|EPI_ISL_447366\|2020-04 | ···················· | ···················· | ···················· | ···················· | ···················· | ···················· |
| C | 2318 | hCoV-19/Israel/2089712/2020\|EPI_ISL_447367\|2020-04 | ···················· | ···················· | ···················· | ···················· | ···················· | ···················· |
| C | 2319 | hCoV-19/Israel/2089697/2020\|EPI_ISL_447369\|2020-04 | ···················· | ···················· | ···················· | ···················· | ···················· | ···················· |
| C | 2320 | hCoV-19/England/OXON-AE2D1/2020\|EPI_ISL_448680\|202 | ···················· | ···················· | ···················· | ···················· | ···················· | ···················· |
| C | 2321 | hCoV-19/England/OXON-AE2FF/2020\|EPI_ISL_448681\|202 | ···················· | ···················· | ···················· | ···················· | ···················· | ···················· |
| C | 2322 | hCoV-19/England/OXON-AE31A/2020\|EPI_ISL_448682\|202 | ···················· | ···················· | ···················· | ···················· | ···················· | ···················· |
| C | 2323 | hCoV-19/England/OXON-AE338/2020\|EPI_ISL_448684\|202 | ···················· | ···················· | ···················· | ···················· | ···················· | ···················· |
| C | 2324 | hCoV-19/England/OXON-AE374/2020\|EPI_ISL_448685\|202 | ···················· | ···················· | ···················· | ···················· | ···················· | ···················· |
| C | 2325 | hCoV-19/England/OXON-AE392/2020\|EPI_ISL_448686\|202 | ···················· | ···················· | ···················· | ···················· | ···················· | ···················· |
| C | 2326 | hCoV-19/England/OXON-AE3B0/2020\|EPI_ISL_448687\|202 | ···················· | ···················· | ···················· | ···················· | ···················· | ···················· |
| C | 2327 | hCoV-19/England/OXON-AE3DE/2020\|EPI_ISL_448688\|202 | ···················· | ···················· | ···················· | ···················· | ···················· | ···················· |
| C | 2328 | hCoV-19/England/OXON-AE3FC/2020\|EPI_ISL_448689\|202 | ···················· | ···················· | ···················· | ···················· | ···················· | ···················· |
| C | 2329 | hCoV-19/England/OXON-AE417/2020\|EPI_ISL_448690\|202 | ···················· | ···················· | ···················· | ···················· | ···················· | ···················· |
| C | 2330 | hCoV-19/England/OXON-AE453/2020\|EPI_ISL_448691\|202 | ···················· | ···················· | ···················· | ···················· | ···················· | ···················· |
| C | 2331 | hCoV-19/England/OXON-AE471/2020\|EPI_ISL_448692\|202 | ···················· | ···················· | ···················· | ···················· | ···················· | ···················· |
| C | 2332 | hCoV-19/England/OXON-AE49F/2020\|EPI_ISL_448694\|202 | ···················· | ···················· | ···················· | ···················· | ···················· | ···················· |
| C | 2333 | hCoV-19/England/OXON-AE4AE/2020\|EPI_ISL_448695\|202 | ···················· | ···················· | ···················· | ···················· | ···················· | ···················· |
| C | 2334 | hCoV-19/England/OXON-AE4BD/2020\|EPI_ISL_448696\|202 | ···················· | ···················· | ···················· | ···················· | ···················· | ···················· |
| C | 2335 | hCoV-19/England/OXON-AE4DB/2020\|EPI_ISL_448697\|202 | ···················· | ···················· | ···················· | ···················· | ···················· | ···················· |
| C | 2336 | hCoV-19/England/OXON-AE4F9/2020\|EPI_ISL_448698\|202 | ···················· | ···················· | ···················· | ···················· | ···················· | ···················· |
| C | 2337 | hCoV-19/England/OXON-AE514/2020\|EPI_ISL_448699\|202 | ···················· | ···················· | ···················· | ···················· | ···················· | ···················· |
| C | 2338 | hCoV-19/England/LIVE-9DFA1/2020\|EPI_ISL_453255\|202 | ···················· | ···················· | ···················· | ···················· | ···················· | ···················· |
| C | 2339 | hCoV-19/Kazakhstan/18243/2020\|EPI_ISL_454500\|2020- | ···················· | ···················· | ···················· | ···················· | ···················· | ···················· |
| C | 2340 | hCoV-19/Kazakhstan/20679/2020\|EPI_ISL_454501\|2020- | ···················· | ···················· | ···················· | ···················· | ···················· | ···················· |
| C | 2341 | hCoV-19/Kazakhstan/21399/2020\|EPI_ISL_454502\|2020- | ···················· | ···················· | ···················· | ···················· | ···················· | ···················· |
| C | 2342 | hCoV-19/Kazakhstan/21927/2020\|EPI_ISL_454503\|2020- | ···················· | ···················· | ···················· | ···················· | ···················· | ···················· |
| C | 2343 | hCoV-19/Kazakhstan/26473/2020\|EPI_ISL_454504\|2020- | ···················· | ···················· | ···················· | ···················· | ···················· | ···················· |
| C | 2344 | hCoV-19/Kazakhstan/26474/2020\|EPI_ISL_454505\|2020- | ···················· | ···················· | ···················· | ···················· | ···················· | ···················· |
| C | 2345 | hCoV-19/Kazakhstan/26478/2020\|EPI_ISL_454506\|2020- | ···················· | ···················· | ···················· | ···················· | ···················· | ···················· |
| C | 2346 | hCoV-19/Kazakhstan/26489/2020\|EPI_ISL_454507\|2020- | ···················· | ···················· | ···················· | ···················· | ···················· | ···················· |
| C | 2347 | hCoV-19/Kazakhstan/26491/2020\|EPI_ISL_454508\|2020- | ···················· | ···················· | ···················· | ···················· | ···················· | ···················· |
| C | 2348 | hCoV-19/Kazakhstan/26497/2020\|EPI_ISL_454509\|2020- | ···················· | ···················· | ···················· | ···················· | ···················· | ···················· |
| C | 2349 | hCoV-19/Kazakhstan/26501/2020\|EPI_ISL_454510\|2020- | ···················· | ···················· | ···················· | ···················· | ···················· | ···················· |
| C | 2350 | hCoV-19/Kazakhstan/26506/2020\|EPI_ISL_454511\|2020- | ···················· | ···················· | ···················· | ···················· | ···················· | ···················· |
| C | 2351 | hCoV-19/Kazakhstan/26548/2020\|EPI_ISL_454512\|2020- | ···················· | ···················· | ···················· | ···················· | ···················· | ···················· |
| C | 2352 | hCoV-19/Kazakhstan/26549/2020\|EPI_ISL_454513\|2020- | ···················· | ···················· | ···················· | ···················· | ···················· | ···················· |
| C | 2353 | hCoV-19/Kazakhstan/26585/2020\|EPI_ISL_454514\|2020- | ···················· | ···················· | ···················· | ···················· | ···················· | ···················· |
| C | 2354 | hCoV-19/Kazakhstan/26617/2020\|EPI_ISL_454515\|2020- | ···················· | ···················· | ···················· | ···················· | ···················· | ···················· |
| C | 2355 | hCoV-19/Kazakhstan/26827/2020\|EPI_ISL_454516\|2020- | ···················· | ···················· | ···················· | ···················· | ···················· | ···················· |
| C | 2356 | hCoV-19/Kazakhstan/26828/2020\|EPI_ISL_454517\|2020- | ···················· | ···················· | ···················· | ···················· | ···················· | ···················· |
| C | 2357 | hCoV-19/Kazakhstan/26829/2020\|EPI_ISL_454518\|2020- | ···················· | ···················· | ···················· | ···················· | ···················· | ···················· |
| C | 2358 | hCoV-19/Kazakhstan/34391/2020\|EPI_ISL_454519\|2020- | ···················· | ···················· | ···················· | ···················· | ···················· | ···················· |
| C | 2359 | hCoV-19/Kazakhstan/38533/2020\|EPI_ISL_454520\|2020- | ···················· | ···················· | ···················· | ···················· | ···················· | ···················· |
| C | 2360 | hCoV-19/India/MH-NIV-11182/2020\|EPI_ISL_454521\|202 | ···················· | ···················· | ···················· | ···················· | ···················· | ···················· |
| C | 2361 | hCoV-19/India/MH-NIV-11676/2020\|EPI_ISL_454522\|202 | ···················· | ···················· | ···················· | ···················· | ···················· | ···················· |
| C | 2366 | hCoV-19/India/MH-NIV-3472/2020\|EPI_ISL_454528\|2020 | ···················· | ···················· | ···················· | ···················· | ···················· | ···················· |
| C | 2368 | hCoV-19/India/MH-NIV-4271/2020\|EPI_ISL_454530\|2020 | ···················· | ···················· | ···················· | ···················· | ···················· | ···················· |
| C | 2371 | hCoV-19/India/MH-NIV-4438/2020\|EPI_ISL_454533\|2020 | ···················· | ···················· | ···················· | ···················· | ···················· | ···················· |
| C | 2372 | hCoV-19/India/MH-NIV-4622/2020\|EPI_ISL_454534\|2020 | ···················· | ···················· | ···················· | ···················· | ···················· | ···················· |
| C | 2375 | hCoV-19/India/MH-NIV-6216/2020\|EPI_ISL_454540\|2020 | ···················· | ···················· | ···················· | ···················· | ···················· | ···················· |
| C | 2376 | hCoV-19/India/MH-NIV-6315/2020\|EPI_ISL_454542\|2020 | ···················· | ···················· | ···················· | ···················· | ···················· | ···················· |
| C | 2378 | hCoV-19/India/MH-NIV-6614/2020\|EPI_ISL_454544\|2020 | ···················· | ···················· | ···················· | ···················· | ···················· | ···················· |
| C | 2379 | hCoV-19/India/MH-NIV-6964/2020\|EPI_ISL_454546\|2020 | ···················· | ···················· | ···················· | ···················· | ···················· | ···················· |
| C | 2380 | hCoV-19/India/MH-NIV-7044/2020\|EPI_ISL_454547\|2020 | ···················· | ···················· | ···················· | ···················· | ···················· | ···················· |
| C | 2381 | hCoV-19/India/MH-NIV-7558/2020\|EPI_ISL_454549\|2020 | ···················· | ···················· | ···················· | ···················· | ···················· | ···················· |
| C | 2385 | hCoV-19/India/MH-NIV-9314/2020\|EPI_ISL_454558\|2020 | ···················· | ···················· | ···················· | ···················· | ···················· | ···················· |
| C | 2387 | hCoV-19/India/MH-NIV-729-3/2020\|EPI_ISL_454561\|202 | ···················· | ···················· | ···················· | ···················· | ···················· | ···················· |
| C | 2388 | hCoV-19/India/MH-NIV-815-3/2020\|EPI_ISL_454562\|202 | ···················· | ···················· | ···················· | ···················· | ···················· | ···················· |
| C | 2389 | hCoV-19/India/MH-NIV-QA-707/2020\|EPI_ISL_454563\|20 | ···················· | ···················· | ···················· | ···················· | ···················· | ···················· |

| | | | | | | | | |
|---|---|---|---|---|---|---|---|---|
| C | 2392 | hCoV-19/Kazakhstan/17920/2020\|EPI_ISL_454577\|2020- | .................... | .................... | .................... | .................... | .................... | .................... |
| C | 2393 | hCoV-19/Croatia/592_Osijek/2020\|EPI_ISL_454578\|202 | .................... | .................... | .................... | .................... | .................... | .................... |
| C | 2394 | hCoV-19/Kazakhstan/18148/2020\|EPI_ISL_454579\|2020- | .................... | .................... | .................... | .................... | .................... | .................... |
| C | 2395 | hCoV-19/Kazakhstan/18287/2020\|EPI_ISL_454580\|2020- | .................... | .................... | .................... | .................... | .................... | .................... |
| C | 2396 | hCoV-19/Croatia/1146_Split/2020\|EPI_ISL_454581\|202 | .................... | .................... | .................... | .................... | .................... | .................... |
| C | 2397 | hCoV-19/Kazakhstan/22001/2020\|EPI_ISL_454582\|2020- | .................... | .................... | .................... | .................... | .................... | .................... |
| C | 2398 | hCoV-19/Croatia/1560_Split/2020\|EPI_ISL_454583\|202 | .................... | .................... | .................... | .................... | .................... | .................... |
| C | 2399 | hCoV-19/Kazakhstan/22044/2020\|EPI_ISL_454584\|2020- | .................... | .................... | .................... | .................... | .................... | .................... |
| C | 2400 | hCoV-19/Kazakhstan/33496/2020\|EPI_ISL_454585\|2020- | .................... | .................... | .................... | .................... | .................... | .................... |
| C | 2401 | hCoV-19/Kazakhstan/22517/2020\|EPI_ISL_454586\|2020- | .................... | .................... | .................... | .................... | .................... | .................... |
| C | 2402 | hCoV-19/Kazakhstan/26475/2020\|EPI_ISL_454587\|2020- | .................... | .................... | .................... | .................... | .................... | .................... |
| C | 2403 | hCoV-19/Croatia/1761_Dubrovnik/2020\|EPI_ISL_454588 | .................... | .................... | .................... | .................... | .................... | .................... |
| C | 2404 | hCoV-19/Kazakhstan/26508/2020\|EPI_ISL_454589\|2020- | .................... | .................... | .................... | .................... | .................... | .................... |
| C | 2405 | hCoV-19/Kazakhstan/26530/2020\|EPI_ISL_454590\|2020- | .................... | .................... | .................... | .................... | .................... | .................... |
| C | 2406 | hCoV-19/Kazakhstan/26545/2020\|EPI_ISL_454591\|2020- | .................... | .................... | .................... | .................... | .................... | .................... |
| C | 2407 | hCoV-19/Croatia/297_Varazdin/2020\|EPI_ISL_454592\|2 | .................... | .................... | .................... | .................... | .................... | .................... |
| C | 2408 | hCoV-19/Kazakhstan/26574/2020\|EPI_ISL_454593\|2020- | .................... | .................... | .................... | .................... | .................... | .................... |
| C | 2409 | hCoV-19/Kazakhstan/26576/2020\|EPI_ISL_454594\|2020- | .................... | .................... | .................... | .................... | .................... | .................... |
| C | 2410 | hCoV-19/Kazakhstan/26577/2020\|EPI_ISL_454596\|2020- | .................... | .................... | .................... | .................... | .................... | .................... |
| C | 2411 | hCoV-19/Kazakhstan/26582/2020\|EPI_ISL_454597\|2020- | .................... | .................... | .................... | .................... | .................... | .................... |
| C | 2412 | hCoV-19/Kazakhstan/26584/2020\|EPI_ISL_454598\|2020- | .................... | .................... | .................... | .................... | .................... | .................... |
| C | 2413 | hCoV-19/Kazakhstan/26562/2020\|EPI_ISL_454599\|2020- | .................... | .................... | .................... | .................... | .................... | .................... |
| C | 2414 | hCoV-19/Sweden/20-50276/2020\|EPI_ISL_455840\|2020-0 | .................... | .................... | .................... | .................... | .................... | .................... |
| C | 2415 | hCoV-19/Sweden/20-50280/2020\|EPI_ISL_455841\|2020-0 | .................... | .................... | .................... | .................... | .................... | .................... |
| C | 2416 | hCoV-19/Sweden/20-08156/2020\|EPI_ISL_455842\|2020-0 | .................... | .................... | .................... | .................... | .................... | .................... |
| C | 2417 | hCoV-19/Sweden/20-08158/2020\|EPI_ISL_455843\|2020-0 | .................... | .................... | .................... | .................... | .................... | .................... |
| C | 2418 | hCoV-19/Sweden/20-08162/2020\|EPI_ISL_455844\|2020-0 | .................... | .................... | .................... | .................... | .................... | .................... |
| C | 2419 | hCoV-19/Sweden/20-08179/2020\|EPI_ISL_455845\|2020-0 | .................... | .................... | .................... | .................... | .................... | .................... |
| C | 2420 | hCoV-19/Sweden/20-08069/2020\|EPI_ISL_455846\|2020-0 | .................... | .................... | .................... | .................... | .................... | .................... |
| C | 2421 | hCoV-19/Sweden/20-08117/2020\|EPI_ISL_455847\|2020-0 | .................... | .................... | .................... | .................... | .................... | .................... |
| C | 2422 | hCoV-19/Sweden/20-50056/2020\|EPI_ISL_455848\|2020-0 | .................... | .................... | .................... | .................... | .................... | .................... |
| C | 2423 | hCoV-19/Sweden/20-50092/2020\|EPI_ISL_455849\|2020-0 | .................... | .................... | .................... | .................... | .................... | .................... |
| C | 2424 | hCoV-19/Sweden/20-50094/2020\|EPI_ISL_455850\|2020-0 | .................... | .................... | .................... | .................... | .................... | .................... |
| C | 2425 | hCoV-19/Sweden/20-50095/2020\|EPI_ISL_455851\|2020-0 | .................... | .................... | .................... | .................... | .................... | .................... |
| C | 2426 | hCoV-19/Sweden/20-50097/2020\|EPI_ISL_455852\|2020-0 | .................... | .................... | .................... | .................... | .................... | .................... |
| C | 2427 | hCoV-19/Sweden/20-50100/2020\|EPI_ISL_455853\|2020-0 | .................... | .................... | .................... | .................... | .................... | .................... |
| C | 2428 | hCoV-19/Sweden/20-50103/2020\|EPI_ISL_455854\|2020-0 | .................... | .................... | .................... | .................... | .................... | .................... |
| C | 2429 | hCoV-19/Sweden/20-50105/2020\|EPI_ISL_455855\|2020-0 | .................... | .................... | .................... | .................... | .................... | .................... |
| C | 2430 | hCoV-19/Sweden/20-50140/2020\|EPI_ISL_455856\|2020-0 | .................... | .................... | .................... | .................... | .................... | .................... |
| C | 2431 | hCoV-19/Sweden/20-50141/2020\|EPI_ISL_455857\|2020-0 | .................... | .................... | .................... | .................... | .................... | .................... |
| C | 2432 | hCoV-19/Sweden/20-50148/2020\|EPI_ISL_455858\|2020-0 | .................... | .................... | .................... | .................... | .................... | .................... |
| C | 2433 | hCoV-19/Sweden/20-50149/2020\|EPI_ISL_455859\|2020-0 | .................... | .................... | .................... | .................... | .................... | .................... |
| C | 2434 | hCoV-19/Sweden/20-50150/2020\|EPI_ISL_455860\|2020-0 | .................... | .................... | .................... | .................... | .................... | .................... |
| C | 2435 | hCoV-19/Sweden/20-50154/2020\|EPI_ISL_455861\|2020-0 | .................... | .................... | .................... | .................... | .................... | .................... |
| C | 2436 | hCoV-19/Sweden/20-50156/2020\|EPI_ISL_455862\|2020-0 | .................... | .................... | .................... | .................... | .................... | .................... |
| C | 2437 | hCoV-19/Sweden/20-50194/2020\|EPI_ISL_455863\|2020-0 | .................... | .................... | .................... | .................... | .................... | .................... |
| C | 2438 | hCoV-19/Sweden/20-50196/2020\|EPI_ISL_455864\|2020-0 | .................... | .................... | .................... | .................... | .................... | .................... |
| C | 2439 | hCoV-19/Sweden/20-50198/2020\|EPI_ISL_455865\|2020-0 | .................... | .................... | .................... | .................... | .................... | .................... |
| C | 2440 | hCoV-19/Sweden/20-50199/2020\|EPI_ISL_455866\|2020-0 | .................... | .................... | .................... | .................... | .................... | .................... |
| C | 2441 | hCoV-19/Sweden/20-50201/2020\|EPI_ISL_455867\|2020-0 | .................... | .................... | .................... | .................... | .................... | .................... |
| C | 2442 | hCoV-19/Sweden/20-50202/2020\|EPI_ISL_455868\|2020-0 | .................... | .................... | .................... | .................... | .................... | .................... |
| C | 2443 | hCoV-19/Sweden/20-50206/2020\|EPI_ISL_455869\|2020-0 | .................... | .................... | .................... | .................... | .................... | .................... |
| C | 2444 | hCoV-19/Sweden/20-50209/2020\|EPI_ISL_455870\|2020-0 | .................... | .................... | .................... | .................... | .................... | .................... |
| C | 2445 | hCoV-19/Sweden/20-50212/2020\|EPI_ISL_455871\|2020-0 | .................... | .................... | .................... | .................... | .................... | .................... |
| C | 2446 | hCoV-19/Sweden/20-50213/2020\|EPI_ISL_455872\|2020-0 | .................... | .................... | .................... | .................... | .................... | .................... |
| C | 2447 | hCoV-19/Sweden/20-50214/2020\|EPI_ISL_455873\|2020-0 | .................... | .................... | .................... | .................... | .................... | .................... |
| C | 2448 | hCoV-19/Sweden/20-50217/2020\|EPI_ISL_455874\|2020-0 | .................... | .................... | .................... | .................... | .................... | .................... |
| C | 2449 | hCoV-19/Sweden/20-50218/2020\|EPI_ISL_455875\|2020-0 | .................... | .................... | .................... | .................... | .................... | .................... |
| C | 2450 | hCoV-19/Sweden/20-50219/2020\|EPI_ISL_455876\|2020-0 | .................... | .................... | .................... | .................... | .................... | .................... |
| C | 2451 | hCoV-19/Sweden/20-50221/2020\|EPI_ISL_455877\|2020-0 | .................... | .................... | .................... | .................... | .................... | .................... |
| C | 2452 | hCoV-19/Sweden/20-50223/2020\|EPI_ISL_455878\|2020-0 | .................... | .................... | .................... | .................... | .................... | .................... |
| C | 2453 | hCoV-19/Sweden/20-50229/2020\|EPI_ISL_455879\|2020-0 | .................... | .................... | .................... | .................... | .................... | .................... |
| C | 2454 | hCoV-19/Sweden/20-50238/2020\|EPI_ISL_455880\|2020-0 | .................... | .................... | .................... | .................... | .................... | .................... |
| C | 2455 | hCoV-19/Sweden/20-50240/2020\|EPI_ISL_455881\|2020-0 | .................... | .................... | .................... | .................... | .................... | .................... |
| C | 2456 | hCoV-19/Sweden/20-50241/2020\|EPI_ISL_455882\|2020-0 | .................... | .................... | .................... | .................... | .................... | .................... |
| C | 2457 | hCoV-19/Sweden/20-50242/2020\|EPI_ISL_455883\|2020-0 | .................... | .................... | .................... | .................... | .................... | .................... |
| C | 2458 | hCoV-19/Sweden/20-50244/2020\|EPI_ISL_455884\|2020-0 | .................... | .................... | .................... | .................... | .................... | .................... |
| C | 2459 | hCoV-19/Sweden/20-50245/2020\|EPI_ISL_455885\|2020-0 | .................... | .................... | .................... | .................... | .................... | .................... |
| C | 2460 | hCoV-19/Sweden/20-50246/2020\|EPI_ISL_455886\|2020-0 | .................... | .................... | .................... | .................... | .................... | .................... |
| C | 2461 | hCoV-19/Sweden/20-50247/2020\|EPI_ISL_455887\|2020-0 | .................... | .................... | .................... | .................... | .................... | .................... |
| C | 2462 | hCoV-19/Sweden/20-50251/2020\|EPI_ISL_455890\|2020-0 | .................... | .................... | .................... | .................... | .................... | .................... |
| C | 2463 | hCoV-19/Sweden/20-50253/2020\|EPI_ISL_455891\|2020-0 | .................... | .................... | .................... | .................... | .................... | .................... |
| C | 2464 | hCoV-19/Sweden/20-50254/2020\|EPI_ISL_455892\|2020-0 | .................... | .................... | .................... | .................... | .................... | .................... |
| C | 2465 | hCoV-19/Sweden/20-50255/2020\|EPI_ISL_455893\|2020-0 | .................... | .................... | .................... | .................... | .................... | .................... |
| C | 2466 | hCoV-19/Sweden/20-50256/2020\|EPI_ISL_455894\|2020-0 | .................... | .................... | .................... | .................... | .................... | .................... |
| C | 2467 | hCoV-19/Sweden/20-50257/2020\|EPI_ISL_455895\|2020-0 | .................... | .................... | .................... | .................... | .................... | .................... |
| C | 2468 | hCoV-19/Sweden/20-50258/2020\|EPI_ISL_455896\|2020-0 | .................... | .................... | .................... | .................... | .................... | .................... |
| C | 2469 | hCoV-19/USA/NC_200468/2020\|EPI_ISL_458004\|2020-04 | .................... | .................... | .................... | .................... | .................... | .................... |

| | | | | | | | | |
|---|---|---|---|---|---|---|---|---|
| C | 2470 | hCoV-19/USA/NY-NYUMC878/2020\|EPI_ISL_458017\|2020-0 | ···················· | ···················· | ···················· | ···················· | ···················· | ···················· |
| C | 2471 | hCoV-19/USA/NY-NYUMC879/2020\|EPI_ISL_458018\|2020-0 | ···················· | ···················· | ···················· | ···················· | ···················· | ···················· |
| C | 2472 | hCoV-19/USA/NY-NYUMC880/2020\|EPI_ISL_458019\|2020-0 | ···················· | ···················· | ···················· | ···················· | ···················· | ···················· |
| C | 2473 | hCoV-19/India/TG-CCMB-L1103/2020\|EPI_ISL_458060\|20 | ···················· | ···················· | ···················· | ···················· | ···················· | ···················· |
| C | 2474 | hCoV-19/India/TG-CCMB-L1108/2020\|EPI_ISL_458061\|20 | ···················· | ···················· | ···················· | ···················· | ···················· | ···················· |
| C | 2475 | hCoV-19/India/TG-CCMB-L299/2020\|EPI_ISL_458062\|202 | ···················· | ···················· | ···················· | ···················· | ···················· | ···················· |
| C | 2476 | hCoV-19/India/TG-CCMB-L301/2020\|EPI_ISL_458063\|202 | ···················· | ···················· | ···················· | ···················· | ···················· | ···················· |
| C | 2477 | hCoV-19/India/TG-CCMB-L988/2020\|EPI_ISL_458064\|202 | ···················· | ···················· | ···················· | ···················· | ···················· | ···················· |
| C | 2478 | hCoV-19/India/TG-CCMB-L989/2020\|EPI_ISL_458065\|202 | ···················· | ···················· | ···················· | ···················· | ···················· | ···················· |
| C | 2479 | hCoV-19/India/TG-CCMB-O1/2020\|EPI_ISL_458066\|2020- | ···················· | ···················· | ···················· | ···················· | ···················· | ···················· |
| C | 2480 | hCoV-19/India/TG-CCMB-O2/2020\|EPI_ISL_458067\|2020- | ···················· | ···················· | ···················· | ···················· | ···················· | ···················· |
| C | 2481 | hCoV-19/India/TG-CCMB-O3/2020\|EPI_ISL_458068\|2020- | ···················· | ···················· | ···················· | ···················· | ···················· | ···················· |
| C | 2482 | hCoV-19/India/TG-CCMB-O4/2020\|EPI_ISL_458069\|2020- | ···················· | ···················· | ···················· | ···················· | ···················· | ···················· |
| C | 2483 | hCoV-19/India/TG-CCMB-K272/2020\|EPI_ISL_458070\|202 | ···················· | ···················· | ···················· | ···················· | ···················· | ···················· |
| C | 2484 | hCoV-19/India/TG-CCMB-J130/2020\|EPI_ISL_458071\|202 | ···················· | ···················· | ···················· | ···················· | ···················· | ···················· |
| C | 2485 | hCoV-19/India/TG-CCMB-J812-P1/2020\|EPI_ISL_458072\| | ···················· | ···················· | ···················· | ···················· | ···················· | ···················· |
| C | 2486 | hCoV-19/India/TG-CCMB-L276-P1/2020\|EPI_ISL_458073\| | ···················· | ···················· | ···················· | ···················· | ···················· | ···················· |
| C | 2487 | hCoV-19/India/TG-CCMB-L301-P1/2020\|EPI_ISL_458074\| | ···················· | ···················· | ···················· | ···················· | ···················· | ···················· |
| C | 2488 | hCoV-19/India/TG-CCMB-O2-P1/2020\|EPI_ISL_458075\|20 | ···················· | ···················· | ···················· | ···················· | ···················· | ···················· |
| C | 2489 | hCoV-19/India/TG-CCMB-L302-P1/2020\|EPI_ISL_458076\| | ···················· | ···················· | ···················· | ···················· | ···················· | ···················· |
| C | 2490 | hCoV-19/India/TG-CCMB-L040-P1/2020\|EPI_ISL_458077\| | ···················· | ···················· | ···················· | ···················· | ···················· | ···················· |
| C | 2491 | hCoV-19/Indonesia/JI-ITD-1238Sp/2020\|EPI_ISL_45807 | ···················· | ···················· | ···················· | ···················· | ···················· | ···················· |
| C | 2492 | hCoV-19/India/TG-CCMB-J300/2020\|EPI_ISL_458080\|202 | ···················· | ···················· | ···················· | ···················· | ···················· | ···················· |
| C | 2493 | hCoV-19/Indonesia/JI-ITD-1273NT/2020\|EPI_ISL_45808 | ···················· | ···················· | ···················· | ···················· | ···················· | ···················· |
| C | 2494 | hCoV-19/Indonesia/JI-ITD-3101NT/2020\|EPI_ISL_45808 | ···················· | ···················· | ···················· | ···················· | ···················· | ···················· |
| C | 2495 | hCoV-19/Italy/SAR-ATS-04039046/2020\|EPI_ISL_458084 | ···················· | ···················· | ···················· | ···················· | ···················· | ···················· |
| C | 2496 | hCoV-19/Italy/SAR-AMVRC-28/2020\|EPI_ISL_458085\|202 | ···················· | ···················· | ···················· | ···················· | ···················· | ···················· |
| C | 2497 | hCoV-19/India/GJ-GBRC131/2020\|EPI_ISL_458086\|2020- | ···················· | ·A·················· | ···················· | ···················· | ···················· | ···A················ |
| C | 2498 | hCoV-19/India/GJ-GBRC132/2020\|EPI_ISL_458087\|2020- | ···················· | ···················· | ···················· | ···················· | ···················· | ···················· |
| C | 2499 | hCoV-19/India/GJ-GBRC133/2020\|EPI_ISL_458088\|2020- | ···················· | ·A·················· | ···················· | ···················· | ···················· | ···A················ |
| C | 2500 | hCoV-19/India/GJ-GBRC134/2020\|EPI_ISL_458089\|2020- | ···················· | ···················· | ···················· | ···················· | ···················· | ···················· |
| C | 2501 | hCoV-19/India/GJ-GBRC135a/2020\|EPI_ISL_458090\|2020 | ···················· | ·A·················· | ···················· | ···················· | ···················· | ···A················ |
| C | 2502 | hCoV-19/India/GJ-GBRC135b/2020\|EPI_ISL_458091\|2020 | ···················· | ·A·················· | ···················· | ···················· | ···················· | ···A················ |
| C | 2503 | hCoV-19/India/GJ-GBRC136a/2020\|EPI_ISL_458092\|2020 | ···················· | ·A·················· | ···················· | ···················· | ···················· | ···A················ |
| C | 2504 | hCoV-19/India/GJ-GBRC136b/2020\|EPI_ISL_458093\|2020 | ···················· | ·A·················· | ···················· | ···················· | ···················· | ···A················ |
| C | 2505 | hCoV-19/India/GJ-GBRC137a/2020\|EPI_ISL_458094\|2020 | ···················· | ·A·················· | ···················· | ···················· | ···················· | ···A················ |
| C | 2506 | hCoV-19/India/GJ-GBRC137b/2020\|EPI_ISL_458095\|2020 | ···················· | ·A·················· | ···················· | ···················· | ···················· | ···A················ |
| C | 2507 | hCoV-19/India/GJ-GBRC138/2020\|EPI_ISL_458096\|2020- | ···················· | ···················· | ···················· | ···················· | ···················· | ···················· |
| C | 2508 | hCoV-19/India/GJ-GBRC139a/2020\|EPI_ISL_458097\|2020 | ···················· | ·A·················· | ···················· | ···················· | ···················· | ···A················ |
| C | 2509 | hCoV-19/India/GJ-GBRC139b/2020\|EPI_ISL_458098\|2020 | ···················· | ·A·················· | ···················· | ···················· | ···················· | ···A················ |
| C | 2510 | hCoV-19/India/GJ-GBRC140/2020\|EPI_ISL_458099\|2020- | ···················· | ·A·················· | ···················· | ···················· | ···················· | ···A················ |
| C | 2511 | hCoV-19/Northern Ireland/NIRE-FA643/2020\|EPI_ISL_4 | ···················· | ···················· | ···················· | ···················· | ···················· | ···················· |
| C | 2512 | hCoV-19/Northern Ireland/NIRE-FC25C/2020\|EPI_ISL_4 | ···················· | ···················· | ···················· | ···················· | ···················· | ···················· |
| C | 2513 | hCoV-19/Northern Ireland/NIRE-FC438/2020\|EPI_ISL_4 | ···················· | ···················· | ···················· | ···················· | ···················· | ···················· |
| C | 2514 | hCoV-19/Northern Ireland/NIRE-FAB17/2020\|EPI_ISL_4 | ···················· | ···················· | ···················· | ···················· | ···················· | ···················· |
| C | 2515 | hCoV-19/Northern Ireland/NIRE-FBFDE/2020\|EPI_ISL_4 | ···················· | ···················· | ···················· | ···················· | ···················· | ···················· |
| C | 2516 | hCoV-19/Northern Ireland/NIRE-FC8D2/2020\|EPI_ISL_4 | ···················· | ···················· | ···················· | ···················· | ···················· | ···················· |
| C | 2517 | hCoV-19/Northern Ireland/NIRE-FBF92/2020\|EPI_ISL_4 | ···················· | ···················· | ···················· | ···················· | ···················· | ···················· |
| C | 2518 | hCoV-19/Northern Ireland/NIRE-FC74E/2020\|EPI_ISL_4 | ···················· | ···················· | ···················· | ···················· | ···················· | ···················· |
| C | 2519 | hCoV-19/Northern Ireland/NIRE-FA6CB/2020\|EPI_ISL_4 | ···················· | ···················· | ···················· | ···················· | ···················· | ···················· |
| C | 2520 | hCoV-19/Northern Ireland/NIRE-FB888/2020\|EPI_ISL_4 | ···················· | ···················· | ···················· | ···················· | ···················· | ···················· |
| C | 2521 | hCoV-19/Northern Ireland/NIRE-FC6D8/2020\|EPI_ISL_4 | ···················· | ···················· | ···················· | ···················· | ···················· | ···················· |
| C | 2522 | hCoV-19/Northern Ireland/NIRE-FAC7E/2020\|EPI_ISL_4 | ···················· | ···················· | ···················· | ···················· | ···················· | ···················· |
| C | 2523 | hCoV-19/Northern Ireland/NIRE-FC68D/2020\|EPI_ISL_4 | ···················· | ···················· | ···················· | ···················· | ···················· | ···················· |
| C | 2524 | hCoV-19/USA/MA-MGH-00425/2020\|EPI_ISL_460313\|2020- | ···················· | ···················· | ···················· | ···················· | ···················· | ···················· |
| C | 2525 | hCoV-19/USA/MA-MGH-00443/2020\|EPI_ISL_460321\|2020- | ···················· | ···················· | ···················· | ···················· | ···················· | ···················· |
| C | 2526 | hCoV-19/USA/MA-MGH-00214/2020\|EPI_ISL_460322\|2020- | ···················· | ···················· | ···················· | ···················· | ···················· | ···················· |
| C | 2527 | hCoV-19/USA/MA-MGH-00721/2020\|EPI_ISL_460323\|2020- | ···················· | ···················· | ···················· | ···················· | ···················· | ···················· |
| C | 2528 | hCoV-19/USA/MA-MGH-00024/2020\|EPI_ISL_460324\|2020- | ···················· | ···················· | ···················· | ···················· | ···················· | ···················· |
| C | 2529 | hCoV-19/USA/MA-MGH-00262/2020\|EPI_ISL_460325\|2020- | ···················· | ···················· | ···················· | ···················· | ···················· | ···················· |
| C | 2530 | hCoV-19/USA/MA-MGH-00097/2020\|EPI_ISL_460326\|2020- | ···················· | ···················· | ···················· | ···················· | ···················· | ···················· |
| C | 2531 | hCoV-19/USA/MA-MGH-00639/2020\|EPI_ISL_460327\|2020- | ···················· | ···················· | ···················· | ···················· | ···················· | ···················· |
| C | 2532 | hCoV-19/USA/MA-MGH-00665/2020\|EPI_ISL_460328\|2020- | ···················· | ···················· | ···················· | ···················· | ···················· | ···················· |
| C | 2533 | hCoV-19/USA/MA-MGH-00188/2020\|EPI_ISL_460329\|2020- | ···················· | ···················· | ···················· | ···················· | ···················· | ···················· |
| C | 2534 | hCoV-19/USA/MA-MGH-00506/2020\|EPI_ISL_460376\|2020- | ···················· | ···················· | ···················· | ···················· | ···················· | ···················· |
| C | 2535 | hCoV-19/USA/MA-MGH-00663/2020\|EPI_ISL_460377\|2020- | ···················· | ···················· | ···················· | ···················· | ···················· | ···················· |
| C | 2536 | hCoV-19/USA/MA-MGH-00256/2020\|EPI_ISL_460378\|2020- | ···················· | ···················· | ···················· | ···················· | ···················· | ···················· |
| C | 2537 | hCoV-19/USA/MA-MGH-00303/2020\|EPI_ISL_460379\|2020- | ···················· | ···················· | ···················· | ···················· | ···················· | ···················· |
| C | 2538 | hCoV-19/USA/MA-MGH-00557/2020\|EPI_ISL_460380\|2020- | ···················· | ···················· | ···················· | ···················· | ···················· | ···················· |
| C | 2540 | hCoV-19/USA/MA-MGH-00600/2020\|EPI_ISL_460382\|2020- | ···················· | ···················· | ···················· | ···················· | ···················· | ···················· |
| C | 2541 | hCoV-19/USA/MA-MGH-00456/2020\|EPI_ISL_460383\|2020- | ···················· | ···················· | ···················· | ···················· | ···················· | ···················· |
| C | 2542 | hCoV-19/USA/MA-MGH-00422/2020\|EPI_ISL_460384\|2020- | ···················· | ···················· | ···················· | ···················· | ···················· | ···················· |
| C | 2543 | hCoV-19/USA/MA-MGH-00468/2020\|EPI_ISL_460385\|2020- | ···················· | ···················· | ···················· | ···················· | ···················· | ···················· |
| C | 2544 | hCoV-19/USA/MA-MGH-00648/2020\|EPI_ISL_460386\|2020- | ···················· | ···················· | ···················· | ···················· | ···················· | ···················· |
| C | 2545 | hCoV-19/USA/MA-MGH-00676/2020\|EPI_ISL_460388\|2020- | ···················· | ···················· | ···················· | ···················· | ···················· | ···················· |
| C | 2546 | hCoV-19/USA/MA-MGH-00234/2020\|EPI_ISL_460389\|2020- | ···················· | ···················· | ···················· | ···················· | ···················· | ···················· |
| C | 2547 | hCoV-19/USA/MA-MGH-00465/2020\|EPI_ISL_460390\|2020- | ···················· | ···················· | ···················· | ···················· | ···················· | ···················· |
| C | 2548 | hCoV-19/USA/MA-MGH-00083/2020\|EPI_ISL_460391\|2020- | ···················· | ···················· | ···················· | ···················· | ···················· | ···················· |

| | | | | | | | | | |
|---|---|---|---|---|---|---|---|---|---|
| C | 2549 hCoV-19/USA/MA-MGH-00479/2020\|EPI_ISL_460392\|2020- | ···················· | ···················· | ···················· | ···················· | ···················· | ···················· | | |
| C | 2550 hCoV-19/USA/MA-MGH-00160/2020\|EPI_ISL_460394\|2020- | ···················· | ···················· | ···················· | ···················· | ···················· | ···················· | | |
| C | 2551 hCoV-19/USA/MA-MGH-00613/2020\|EPI_ISL_460396\|2020- | ···················· | ···················· | ···················· | ···················· | ···················· | ···················· | | |
| C | 2552 hCoV-19/USA/MA-MGH-00424/2020\|EPI_ISL_460397\|2020- | ···················· | ···················· | ···················· | ···················· | ···················· | ···················· | | |
| C | 2553 hCoV-19/USA/MA-MGH-00510/2020\|EPI_ISL_460398\|2020- | ···················· | ···················· | ···················· | ···················· | ···················· | ···················· | | |
| C | 2554 hCoV-19/USA/MA-MGH-00576/2020\|EPI_ISL_460399\|2020- | ···················· | ···················· | ···················· | ···················· | ···················· | ···················· | | |
| C | 2555 hCoV-19/Scotland/CVR3068/2020\|EPI_ISL_461652\|2020- | ···················· | ···················· | ···················· | ···················· | ···················· | ···················· | | |
| C | 2556 hCoV-19/Scotland/CVR3071/2020\|EPI_ISL_461655\|2020- | ···················· | ···················· | ···················· | ···················· | ···················· | ···················· | | |
| C | 2557 hCoV-19/Scotland/CVR3072/2020\|EPI_ISL_461656\|2020- | ···················· | ···················· | ···················· | ···················· | ···················· | ···················· | | |
| C | 2558 hCoV-19/Scotland/CVR3073/2020\|EPI_ISL_461657\|2020- | ···················· | ···················· | ···················· | ···················· | ···················· | ···················· | | |
| C | 2559 hCoV-19/Scotland/CVR3075/2020\|EPI_ISL_461659\|2020- | ···················· | ···················· | ···················· | ···················· | ···················· | ···················· | | |
| C | 2561 hCoV-19/Israel/51140028/2020\|EPI_ISL_447328\|2020-0 | ···················· | ···················· | ···················· | ···················· | ···················· | ···················· | | |
| C | 2562 hCoV-19/Israel/51137031/2020\|EPI_ISL_447330\|2020-0 | ···················· | ···················· | ···················· | ···················· | ···················· | ···················· | | |
| C | 2563 hCoV-19/Israel/2123863/2020\|EPI_ISL_447331\|2020-04 | ···················· | ···················· | ···················· | ···················· | ···················· | ···················· | | |
| C | 2564 hCoV-19/Israel/2123853/2020\|EPI_ISL_447332\|2020-04 | ···················· | ···················· | ···················· | ···················· | ···················· | ···················· | | |
| C | 2565 hCoV-19/Israel/2115990/2020\|EPI_ISL_447334\|2020-04 | ···················· | ···················· | ···················· | ···················· | ···A················ | ···················· | | |
| C | 2566 hCoV-19/Israel/2115968/2020\|EPI_ISL_447337\|2020-04 | ···················· | ···················· | ···················· | ···················· | ···················· | ···················· | | |
| C | 2567 hCoV-19/Israel/2115964/2020\|EPI_ISL_447338\|2020-04 | ···················· | ···················· | ···················· | ···················· | ···················· | ···················· | | |
| C | 2568 hCoV-19/Israel/2115701/2020\|EPI_ISL_447339\|2020-04 | ···················· | ···················· | ···················· | ···················· | ···················· | ···················· | | |
| C | 2569 hCoV-19/Israel/2113678/2020\|EPI_ISL_447340\|2020-04 | ···················· | ···················· | ···················· | ···················· | ···················· | ···················· | | |
| C | 2570 hCoV-19/Israel/2113603/2020\|EPI_ISL_447341\|2020-04 | ···················· | ···················· | ···················· | ···················· | ···················· | ···················· | | |
| C | 2571 hCoV-19/Israel/2113601/2020\|EPI_ISL_447342\|2020-04 | ···················· | ···················· | ···················· | ···················· | ···················· | ···················· | | |
| C | 2572 hCoV-19/Israel/2113256/2020\|EPI_ISL_447343\|2020-04 | ···················· | ···················· | ···················· | ···················· | ···N················ | ···················· | | |
| C | 2574 hCoV-19/Israel/2113178/2020\|EPI_ISL_447345\|2020-04 | ···················· | ···················· | ···················· | ···················· | ···················· | ···················· | | |
| C | 2575 hCoV-19/Israel/2113174/2020\|EPI_ISL_447346\|2020-04 | ···················· | ···················· | ···················· | ···················· | ···················· | ···················· | | |
| C | 2576 hCoV-19/Israel/2113173/2020\|EPI_ISL_447347\|2020-04 | ···················· | ···················· | ···················· | ···················· | ···················· | ···················· | | |
| C | 2577 hCoV-19/Israel/2113161/2020\|EPI_ISL_447348\|2020-04 | ···················· | ···················· | ···················· | ···················· | ···················· | ···················· | | |
| C | 2578 hCoV-19/Israel/2113155/2020\|EPI_ISL_447349\|2020-04 | ···················· | ···················· | ···················· | ···················· | ···················· | ···················· | | |
| C | 2579 hCoV-19/Israel/2089383/2020\|EPI_ISL_447370\|2020-04 | ···················· | ···················· | ···················· | ···················· | ···················· | ···················· | | |
| C | 2580 hCoV-19/Israel/2089375/2020\|EPI_ISL_447372\|2020-04 | ···················· | ···················· | ···················· | ···················· | ···················· | ···················· | | |
| C | 2581 hCoV-19/Israel/2089366/2020\|EPI_ISL_447374\|2020-04 | ···················· | ···················· | ···················· | ···················· | ···················· | ···················· | | |
| C | 2582 hCoV-19/Israel/2086045/2020\|EPI_ISL_447375\|2020-04 | ···················· | ···················· | ···················· | ···················· | ···················· | ···················· | | |
| C | 2583 hCoV-19/Israel/2086012/2020\|EPI_ISL_447379\|2020-04 | ···················· | ···················· | ···················· | ···················· | ···················· | ···················· | | |
| C | 2584 hCoV-19/Israel/2086008/2020\|EPI_ISL_447380\|2020-04 | ···················· | ···················· | ···················· | ···················· | ···················· | ···················· | | |
| C | 2585 hCoV-19/Israel/2086004/2020\|EPI_ISL_447381\|2020-04 | ···················· | ···················· | ···················· | ···················· | ···················· | ···················· | | |
| C | 2586 hCoV-19/Israel/2086001/2020\|EPI_ISL_447382\|2020-04 | ···················· | ···················· | ···················· | ···················· | ···················· | ···················· | | |
| C | 2587 hCoV-19/Israel/2047883/2020\|EPI_ISL_447384\|2020-03 | ···················· | ···················· | ···················· | ···················· | ···················· | ········N···N···N··· | 1 | 1 |
| C | 2588 hCoV-19/Israel/2047772/2020\|EPI_ISL_447385\|2020-03 | ···················· | ···················· | ···················· | ···················· | ···················· | ···N················ | | |
| C | 2589 hCoV-19/Israel/2047749/2020\|EPI_ISL_447386\|2020-03 | ···················· | ···················· | ···················· | ···················· | ···················· | ········N····N······ | | |
| C | 2590 hCoV-19/Israel/2047738/2020\|EPI_ISL_447387\|2020-03 | ···················· | ···················· | ···················· | ···················· | ···················· | ···················· | | |
| C | 2591 hCoV-19/Israel/2047604/2020\|EPI_ISL_447388\|2020-03 | ···················· | ···················· | ···················· | ···················· | ···················· | ···N········NN······ | | 1 |
| C | 2592 hCoV-19/Israel/2047586/2020\|EPI_ISL_447389\|2020-03 | ···················· | ···················· | ···················· | ···················· | ···················· | ···················· | | |
| C | 2593 hCoV-19/Israel/2047418/2020\|EPI_ISL_447391\|2020-03 | ···················· | ···················· | ···················· | ···················· | ···················· | ···················· | | |
| C | 2594 hCoV-19/Israel/2047364/2020\|EPI_ISL_447393\|2020-03 | ···················· | ···················· | ···················· | ···················· | ···················· | ···················· | | |
| C | 2595 hCoV-19/Israel/2047189/2020\|EPI_ISL_447394\|2020-03 | ···················· | ···················· | ···················· | ···················· | ···················· | ···················· | | |
| C | 2596 hCoV-19/Israel/2047188/2020\|EPI_ISL_447395\|2020-03 | ···················· | ···················· | ···················· | ···················· | ···················· | ···················· | | |
| C | 2597 hCoV-19/Israel/2047145/2020\|EPI_ISL_447396\|2020-03 | ···················· | ···················· | ···················· | ···················· | ···················· | ···················· | | |
| C | 2598 hCoV-19/Israel/2047016/2020\|EPI_ISL_447397\|2020-03 | ···················· | ···················· | ···················· | ···················· | ···················· | ···················· | | |
| C | 2599 hCoV-19/Israel/2047004/2020\|EPI_ISL_447399\|2020-03 | ···················· | ···················· | ···················· | ···················· | ···················· | ···················· | | |
| C | 2600 hCoV-19/England/OXON-AD876/2020\|EPI_ISL_448600\|202 | ···················· | ···················· | ···················· | ···················· | ···················· | ···················· | | |
| C | 2601 hCoV-19/England/OXON-AD894/2020\|EPI_ISL_448601\|202 | ···················· | ···················· | ···················· | ···················· | ···················· | ···················· | | |
| C | 2602 hCoV-19/England/OXON-AD8D0/2020\|EPI_ISL_448602\|202 | ···················· | ···················· | ···················· | ···················· | ···················· | ···················· | | |
| C | 2603 hCoV-19/England/OXON-AD8FE/2020\|EPI_ISL_448603\|202 | ···················· | ···················· | ···················· | ···················· | ···················· | ···················· | | |
| C | 2604 hCoV-19/England/OXON-AE07D/2020\|EPI_ISL_448659\|202 | ···················· | ···················· | ···················· | ···················· | ···················· | ···················· | | |
| C | 2605 hCoV-19/England/OXON-AE09B/2020\|EPI_ISL_448660\|202 | ···················· | ···················· | ···················· | ···················· | ···················· | ···················· | | |
| C | 2606 hCoV-19/England/OXON-AE0B9/2020\|EPI_ISL_448661\|202 | ···················· | ···················· | ···················· | ···················· | ···················· | ···················· | | |
| C | 2607 hCoV-19/England/OXON-AE0D7/2020\|EPI_ISL_448662\|202 | ···················· | ···················· | ···················· | ···················· | ···················· | ···················· | | |
| C | 2608 hCoV-19/England/OXON-AE0F5/2020\|EPI_ISL_448663\|202 | ···················· | ···················· | ···················· | ···················· | ···················· | ···················· | | |
| C | 2609 hCoV-19/England/OXON-AE110/2020\|EPI_ISL_448664\|202 | ···················· | ···················· | ···················· | ···················· | ···················· | ···················· | | |
| C | 2610 hCoV-19/England/OXON-AE15C/2020\|EPI_ISL_448666\|202 | ···················· | ···················· | ···················· | ···················· | ···················· | ···················· | | |
| C | 2611 hCoV-19/England/OXON-AE17A/2020\|EPI_ISL_448667\|202 | ···················· | ···················· | ···················· | ···················· | ···················· | ···················· | | |
| C | 2612 hCoV-19/England/OXON-AE189/2020\|EPI_ISL_448668\|202 | ···················· | ···················· | ···················· | ···················· | ···················· | ···················· | | |
| C | 2613 hCoV-19/England/OXON-AE198/2020\|EPI_ISL_448669\|202 | ···················· | ···················· | ···················· | ···················· | ···················· | ···················· | | |
| C | 2614 hCoV-19/England/OXON-AE1B6/2020\|EPI_ISL_448670\|202 | ···················· | ···················· | ···················· | ···················· | ···················· | ···················· | | |
| C | 2615 hCoV-19/England/OXON-AE1D4/2020\|EPI_ISL_448671\|202 | ···················· | ···················· | ···················· | ···················· | ···················· | ···················· | | |
| C | 2616 hCoV-19/England/OXON-AE1F2/2020\|EPI_ISL_448672\|202 | ···················· | ···················· | ···················· | ···················· | ···················· | ···················· | | |
| C | 2617 hCoV-19/England/OXON-AE21D/2020\|EPI_ISL_448673\|202 | ···················· | ···················· | ···················· | ···················· | ···················· | ···················· | | |
| C | 2618 hCoV-19/England/OXON-AE22C/2020\|EPI_ISL_448674\|202 | ···················· | ···················· | ···················· | ···················· | ···················· | ···················· | | |
| C | 2619 hCoV-19/England/OXON-AE23B/2020\|EPI_ISL_448675\|202 | ···················· | ···················· | ···················· | ···················· | ···················· | ···················· | | |
| C | 2620 hCoV-19/England/OXON-AE259/2020\|EPI_ISL_448676\|202 | ···················· | ···················· | ···················· | ···················· | ···················· | ···················· | | |
| C | 2621 hCoV-19/England/OXON-AE295/2020\|EPI_ISL_448678\|202 | ···················· | ···················· | ···················· | ···················· | ···················· | ···················· | | |
| C | 2622 hCoV-19/England/OXON-AE2B3/2020\|EPI_ISL_448679\|202 | ···················· | ···················· | ···················· | ···················· | ···················· | ···················· | | |
| C | 2623 hCoV-19/USA/WA-S755/2020\|EPI_ISL_449925\|2020-04-14 | ···················· | ···················· | ···················· | ···················· | ···················· | ···················· | | |
| C | 2624 hCoV-19/USA/WA-S758/2020\|EPI_ISL_449928\|2020-04-13 | ···················· | ···················· | ···················· | ···················· | ···················· | ···················· | | |
| C | 2625 hCoV-19/USA/WA-S759/2020\|EPI_ISL_449929\|2020-04-14 | ···················· | ···················· | ···················· | ···················· | ···················· | ···················· | | |
| C | 2626 hCoV-19/USA/WA-S760/2020\|EPI_ISL_449930\|2020-04-14 | ···················· | ···················· | ···················· | ···················· | ···················· | ···················· | | |
| C | 2627 hCoV-19/USA/WA-S761/2020\|EPI_ISL_449931\|2020-04-14 | ···················· | ···················· | ···················· | ···················· | ···················· | ···················· | | |
| C | 2628 hCoV-19/USA/WA-S762/2020\|EPI_ISL_449932\|2020-04-14 | ···················· | ···················· | ···················· | ···················· | ···················· | ···················· | | |

| | | | | | | | | |
|---|---|---|---|---|---|---|---|---|
| C | 2629 hCoV-19/USA/WA-S763/2020\|EPI_ISL_449933\|2020-04-14 | ···················· | ···················· | ···················· | ···················· | ···················· | ···················· | |
| C | 2630 hCoV-19/USA/WA-S764/2020\|EPI_ISL_449934\|2020-04-14 | ···················· | ···················· | ···················· | ···················· | ···················· | ···················· | |
| C | 2631 hCoV-19/USA/WA-S820/2020\|EPI_ISL_449990\|2020-04-15 | ···················· | ···················· | ···················· | ···················· | ···················· | ···················· | |
| C | 2632 hCoV-19/USA/WA-S821/2020\|EPI_ISL_449991\|2020-04-16 | ···················· | ···················· | ···················· | ···················· | ···················· | ···················· | |
| C | 2633 hCoV-19/USA/WA-S822/2020\|EPI_ISL_449992\|2020-04-16 | ···················· | ···················· | ···················· | ···················· | ···················· | ···················· | |
| C | 2634 hCoV-19/USA/WA-S824/2020\|EPI_ISL_449994\|2020-04-17 | ···················· | ···················· | ···················· | ···················· | ···················· | ···················· | |
| C | 2635 hCoV-19/USA/WA-S825/2020\|EPI_ISL_449995\|2020-04-16 | ···················· | ···················· | ···················· | ···················· | ···················· | ···················· | |
| C | 2636 hCoV-19/USA/WA-S826/2020\|EPI_ISL_449996\|2020-04-16 | ···················· | ···················· | ···················· | ···················· | ···················· | ···················· | |
| C | 2637 hCoV-19/USA/WA-S827/2020\|EPI_ISL_449997\|2020-04-16 | ···················· | ···················· | ···················· | ···················· | ···················· | ···················· | |
| C | 2638 hCoV-19/USA/WA-S828/2020\|EPI_ISL_449998\|2020-04-16 | ···················· | ···················· | ···················· | ···················· | ···················· | ···················· | |
| C | 2639 hCoV-19/USA/WA-S829/2020\|EPI_ISL_449999\|2020-04-14 | ···················· | ···················· | ···················· | ···················· | ···················· | ···················· | |
| C | 2640 hCoV-19/Kenya/D6/2020\|EPI_ISL_458000\|2020-03-17 | ···················· | ···················· | ···················· | ···················· | ···················· | ···················· | |
| C | 2641 hCoV-19/USA/NC_200460/2020\|EPI_ISL_458002\|2020-04 | ···················· | ···················· | ···················· | ···················· | ···················· | ···················· | |
| C | 2642 hCoV-19/USA/NC_200467/2020\|EPI_ISL_458003\|2020-04 | ···················· | ···················· | ···················· | ···················· | ···················· | ···················· | |
| C | 2643 hCoV-19/USA/NC_200486/2020\|EPI_ISL_458005\|2020-04 | ···················· | ···················· | ···················· | ···················· | ···················· | ···················· | |
| C | 2644 hCoV-19/USA/NC_200488/2020\|EPI_ISL_458006\|2020-04 | ···················· | ···················· | ···················· | ···················· | ···················· | ···················· | |
| C | 2645 hCoV-19/USA/NC_200222/2020\|EPI_ISL_458007\|2020-03 | ···················· | ···················· | ···················· | ···················· | ···················· | ···················· | |
| C | 2646 hCoV-19/USA/NY-NYUMC881/2020\|EPI_ISL_458020\|2020-0 | ···················· | ···················· | ···················· | ···················· | ···················· | ···················· | |
| C | 2647 hCoV-19/USA/NY-NYUMC882/2020\|EPI_ISL_458021\|2020-0 | ···················· | ·············A······ | ···················· | ···················· | ···················· | ···················· | |
| C | 2649 hCoV-19/Taiwan/TSGH-38/2020\|EPI_ISL_458029\|2020-03 | ···················· | ···················· | ···················· | ···················· | ···················· | ···················· | |
| C | 2650 hCoV-19/India/TN-CCMB-C12/2020\|EPI_ISL_458030\|2020 | ···················· | ···················· | ···················· | ···················· | ···················· | ···················· | |
| C | 2651 hCoV-19/India/TN-CCMB-C6/2020\|EPI_ISL_458031\|2020- | ···················· | ···················· | ···················· | ···················· | ···················· | ···················· | |
| C | 2652 hCoV-19/India/TN-CCMB-C8/2020\|EPI_ISL_458032\|2020- | ···················· | ···················· | ···················· | ···················· | ···················· | ···················· | |
| C | 2653 hCoV-19/India/TN-CCMB-C5/2020\|EPI_ISL_458033\|2020- | ···················· | ···················· | ···················· | ···················· | ···················· | ···················· | |
| C | 2654 hCoV-19/India/TN-CCMB-C11/2020\|EPI_ISL_458034\|2020 | ···················· | ···················· | ···················· | ···················· | ···················· | ···················· | |
| C | 2655 hCoV-19/India/TN-CCMB-C13/2020\|EPI_ISL_458035\|2020 | ···················· | ···················· | ···················· | ···················· | ···················· | ···················· | |
| C | 2656 hCoV-19/India/TN-CCMB-C10/2020\|EPI_ISL_458036\|2020 | ···················· | ···················· | ···················· | ···················· | ···················· | ···················· | |
| C | 2657 hCoV-19/India/TN-CCMB-C7/2020\|EPI_ISL_458037\|2020- | ···················· | ···················· | ···················· | ···················· | ···················· | ···················· | |
| C | 2658 hCoV-19/India/TN-CCMB-C9/2020\|EPI_ISL_458038\|2020- | ···················· | ···················· | ···················· | ···················· | ···················· | ···················· | |
| C | 2659 hCoV-19/India/TN-CCMB-C16/2020\|EPI_ISL_458039\|2020 | ···················· | ···················· | ···················· | ···················· | ···················· | ···················· | |
| C | 2660 hCoV-19/India/TN-CCMB-C19/2020\|EPI_ISL_458040\|2020 | ···················· | ···················· | ···················· | ···················· | ···················· | ···················· | |
| C | 2661 hCoV-19/India/TN-CCMB-C15/2020\|EPI_ISL_458041\|2020 | ···················· | ···················· | ···················· | ···················· | ···················· | ···················· | |
| C | 2662 hCoV-19/India/TN-CCMB-C18/2020\|EPI_ISL_458042\|2020 | ···················· | ···················· | ···················· | ···················· | ···················· | ···················· | |
| C | 2663 hCoV-19/India/TN-CCMB-C14/2020\|EPI_ISL_458043\|2020 | ···················· | ···················· | ···················· | ···················· | ···················· | ···················· | |
| C | 2664 hCoV-19/India/TN-CCMB-C17/2020\|EPI_ISL_458044\|2020 | ···················· | ···················· | ···················· | ···················· | ···················· | ···················· | |
| C | 2665 hCoV-19/India/TG-CCMB-L1017/2020\|EPI_ISL_458045\|20 | ···················· | ···················· | ···················· | ···················· | ···················· | ···················· | |
| C | 2666 hCoV-19/India/TG-CCMB-L1021/2020\|EPI_ISL_458046\|20 | ···················· | ···················· | ···················· | ···················· | ···················· | ···················· | |
| C | 2667 hCoV-19/India/TG-CCMB-L1022/2020\|EPI_ISL_458047\|20 | ···················· | ···················· | ···················· | ···················· | ···················· | ···················· | |
| C | 2668 hCoV-19/India/TG-CCMB-L1024/2020\|EPI_ISL_458048\|20 | ···················· | ···················· | ···················· | ···················· | ···················· | ···················· | |
| C | 2669 hCoV-19/India/TG-CCMB-L1025/2020\|EPI_ISL_458049\|20 | ···················· | ···················· | ···················· | ···················· | ···················· | ···················· | |
| C | 2670 hCoV-19/India/TG-CCMB-L1026/2020\|EPI_ISL_458050\|20 | ···················· | ···················· | ···················· | ···················· | ···················· | ···················· | |
| C | 2671 hCoV-19/India/TG-CCMB-L1062/2020\|EPI_ISL_458051\|20 | ···················· | ···················· | ···················· | ···················· | ···················· | ···················· | |
| C | 2672 hCoV-19/India/TG-CCMB-L1063/2020\|EPI_ISL_458052\|20 | ···················· | ···················· | ···················· | ···················· | ···················· | ···················· | |
| C | 2673 hCoV-19/India/TG-CCMB-L1064/2020\|EPI_ISL_458053\|20 | ···················· | ···················· | ···················· | ···················· | ···················· | ···················· | |
| C | 2674 hCoV-19/India/TG-CCMB-L1073/2020\|EPI_ISL_458054\|20 | ···················· | ···················· | ···················· | ···················· | ···················· | ···················· | |
| C | 2675 hCoV-19/India/TG-CCMB-L1082/2020\|EPI_ISL_458055\|20 | ···················· | ···················· | ···················· | ···················· | ···················· | ···················· | |
| C | 2676 hCoV-19/India/TG-CCMB-L1085/2020\|EPI_ISL_458056\|20 | ···················· | ···················· | ···················· | ···················· | ···················· | ···················· | |
| C | 2677 hCoV-19/India/TG-CCMB-L1089/2020\|EPI_ISL_458057\|20 | ···················· | ···················· | ···················· | ···················· | ···················· | ···················· | |
| C | 2678 hCoV-19/India/TG-CCMB-L1090/2020\|EPI_ISL_458058\|20 | ···················· | ···················· | ···················· | ···················· | ···················· | ···················· | |
| C | 2679 hCoV-19/India/TG-CCMB-L1100/2020\|EPI_ISL_458059\|20 | ···················· | ···················· | ···················· | ···················· | ···················· | ···················· | |
| C | 2680 hCoV-19/England/CAMB-1AF099/2020\|EPI_ISL_459309\|20 | ···················· | ···················· | ···················· | ···················· | ···················· | ···················· | |
| C | 2681 hCoV-19/England/CAMB-1AF6C4/2020\|EPI_ISL_459310\|20 | ···················· | ···················· | ···················· | ···················· | ···················· | ···················· | |
| C | 2682 hCoV-19/England/CAMB-1AF1F0/2020\|EPI_ISL_459313\|20 | ···················· | ···················· | ···················· | ···················· | ···················· | ···················· | |
| C | 2683 hCoV-19/England/CAMB-1B0217/2020\|EPI_ISL_459315\|20 | ···················· | ···················· | ···················· | ···················· | ···················· | ···················· | |
| C | 2684 hCoV-19/England/CAMB-1AF952/2020\|EPI_ISL_459316\|20 | ···················· | ···················· | ···················· | ···················· | ···················· | ···················· | |
| C | 2685 hCoV-19/England/CAMB-1AF63D/2020\|EPI_ISL_459317\|20 | ···················· | ···················· | ···················· | ···················· | ···················· | ···················· | |
| C | 2686 hCoV-19/England/CAMB-1AE8FC/2020\|EPI_ISL_459320\|20 | ···················· | ···················· | ···················· | ···················· | ···················· | ···················· | |
| C | 2687 hCoV-19/England/CAMB-1AEFD9/2020\|EPI_ISL_459321\|20 | ···················· | ···················· | ···················· | ···················· | ···················· | ···················· | |
| C | 2688 hCoV-19/Northern Ireland/NIRE-FACBA/2020\|EPI_ISL_4 | ···················· | ···················· | ···················· | ···················· | ···················· | ···················· | |
| C | 2689 hCoV-19/Northern Ireland/NIRE-FA6E9/2020\|EPI_ISL_4 | ···················· | ···················· | ···················· | ···················· | ···················· | ···················· | |
| C | 2690 hCoV-19/Northern Ireland/NIRE-FA625/2020\|EPI_ISL_4 | ···················· | ···················· | ···················· | ···················· | ···················· | ···················· | |
| C | 2691 hCoV-19/Northern Ireland/NIRE-FBA64/2020\|EPI_ISL_4 | ···················· | ···················· | ···················· | ···················· | ···················· | ···················· | |
| C | 2692 hCoV-19/Northern Ireland/NIRE-FC993/2020\|EPI_ISL_4 | ···················· | ···················· | ···················· | ···················· | ···················· | ···················· | |
| C | 2693 hCoV-19/Northern Ireland/NIRE-FC22F/2020\|EPI_ISL_4 | ···················· | ···················A | ···················· | ···················· | ···················· | ···················· | 1 |
| C | 2694 hCoV-19/Northern Ireland/NIRE-FC2C5/2020\|EPI_ISL_4 | ···················· | ···················· | ···················· | ···················· | ···················· | ···················· | |
| C | 2695 hCoV-19/Northern Ireland/NIRE-FA7AA/2020\|EPI_ISL_4 | ···················· | ···················· | ···················· | ···················· | ···················· | ···················· | |
| C | 2696 hCoV-19/Northern Ireland/NIRE-FC948/2020\|EPI_ISL_4 | ···················· | ···················· | ···················· | ···················· | ···················· | ···················· | |
| C | 2697 hCoV-19/Northern Ireland/NIRE-FC7C6/2020\|EPI_ISL_4 | ···················· | ···················· | ···················· | ···················· | ···················· | ···················· | |
| C | 2698 hCoV-19/Northern Ireland/NIRE-FC359/2020\|EPI_ISL_4 | ···················· | ···················· | ···················· | ···················· | ···················· | ···················· | |
| C | 2699 hCoV-19/Northern Ireland/NIRE-FC91B/2020\|EPI_ISL_4 | ···················· | ···················· | ···················· | ···················· | ···················· | ···················· | |
| C | 2700 hCoV-19/Northern Ireland/NIRE-FA661/2020\|EPI_ISL_4 | ···················· | ···················· | ···················· | ···················· | ···················· | ···················· | |
| C | 2701 hCoV-19/Northern Ireland/NIRE-FBD7A/2020\|EPI_ISL_4 | ···················· | ···················· | ···················· | ···················· | ···················· | ···················· | |
| C | 2702 hCoV-19/Northern Ireland/NIRE-FC939/2020\|EPI_ISL_4 | ···················· | ···················· | ···················· | ···················· | ···················· | ···················· | |
| C | 2703 hCoV-19/Northern Ireland/NIRE-FC702/2020\|EPI_ISL_4 | ···················· | ···················· | ···················· | ···················· | ···················· | ···················· | |
| C | 2704 hCoV-19/Northern Ireland/NIRE-FC8A5/2020\|EPI_ISL_4 | ···················· | ···················· | ···················· | ···················· | ···················· | ···················· | |
| C | 2705 hCoV-19/Northern Ireland/NIRE-FC9B1/2020\|EPI_ISL_4 | ···················· | ···················· | ···················· | ···················· | ···················· | ···················· | |
| C | 2706 hCoV-19/Northern Ireland/NIRE-FBF0B/2020\|EPI_ISL_4 | ···················· | ···················· | ···················· | ···················· | ···················· | ···················· | |
| C | 2707 hCoV-19/Northern Ireland/NIRE-FC16E/2020\|EPI_ISL_4 | ···················· | ···················· | ···················· | ···················· | ···················· | ···················· | |

C 2708 hCoV-19/Northern Ireland/NIRE-FA76E/2020|EPI_ISL_4 ···················· ···················· ····················· ···················· ···················· ····················
C 2709 hCoV-19/Northern Ireland/NIRE-FC69C/2020|EPI_ISL_4 ···················· ···················· ····················· ···················· ···················· ····················
C 2710 hCoV-19/Northern Ireland/NIRE-FC7B7/2020|EPI_ISL_4 ···················· ···················· ····················· ···················· ···················· ····················
C 2711 hCoV-19/Northern Ireland/NIRE-FC799/2020|EPI_ISL_4 ···················· ···················· ····················· ···················· ···················· ····················
C 2712 hCoV-19/Northern Ireland/NIRE-FBA28/2020|EPI_ISL_4 ···················· ···················· ····················· ···················· ···················· ····················
C 2713 hCoV-19/Northern Ireland/NIRE-FA801/2020|EPI_ISL_4 ···················· ···················· ····················· ···················· ···················· ····················
C 2714 hCoV-19/Northern Ireland/NIRE-FC984/2020|EPI_ISL_4 ···················· ···················· ····················· ···················· ···················· ····················
C 2715 hCoV-19/Northern Ireland/NIRE-FC4CF/2020|EPI_ISL_4 ···················· ···················· ····················· ···················· ···················· ····················
C 2716 hCoV-19/USA/MA-MGH-00419/2020|EPI_ISL_460300|2020- ···················· ···················· ····················· ···················· ···················· ····················
C 2717 hCoV-19/USA/MA-MGH-00159/2020|EPI_ISL_460301|2020- ···················· ···················· ····················· ···················· ···················· ····················
C 2718 hCoV-19/USA/MA-MGH-00017/2020|EPI_ISL_460302|2020- ···················· ···················· ····················· ···················· ···················· ····················
C 2719 hCoV-19/USA/MA-MGH-00192/2020|EPI_ISL_460304|2020- ···················· ···················· ····················· ···················· ···················· ····················
C 2720 hCoV-19/USA/MA-MGH-00248/2020|EPI_ISL_460305|2020- ···················· ···················· ····················· ···················· ···················· ····················
C 2721 hCoV-19/USA/MA-MGH-00489/2020|EPI_ISL_460306|2020- ···················· ···················· ····················· ···················· ···················· ····················
C 2722 hCoV-19/USA/MA-MGH-00664/2020|EPI_ISL_460307|2020- ···················· ···················· ····················· ···················· ···················· ····················
C 2723 hCoV-19/USA/MA-MGH-00447/2020|EPI_ISL_460308|2020- ···················· ···················· ····················· ···················· ···················· ····················
C 2724 hCoV-19/USA/MA-MGH-00227/2020|EPI_ISL_460309|2020- ···················· ···················· ····················· ···················· ···················· ····················
C 2725 hCoV-19/USA/MA-MGH-00062/2020|EPI_ISL_460310|2020- ···················· ···················· ····················· ···················· ···················· ····················
C 2726 hCoV-19/USA/MA-MGH-00003/2020|EPI_ISL_460311|2020- ···················· ···················· ····················· ···················· ···················· ····················
C 2727 hCoV-19/USA/MA-MGH-00571/2020|EPI_ISL_460312|2020- ···················· ···················· ····················· ···················· ···················· ····················
C 2728 hCoV-19/USA/MA-MGH-00598/2020|EPI_ISL_460314|2020- ···················· ···················· ····················· ···················· ···················· ····················
C 2729 hCoV-19/USA/MA-MGH-00718/2020|EPI_ISL_460315|2020- ···················· ···················· ····················· ···················· ···················· ····················
C 2730 hCoV-19/USA/MA-MGH-00134/2020|EPI_ISL_460316|2020- ···················· ···················· ····················· ···················· ···················· ····················
C 2731 hCoV-19/USA/MA-MGH-00592/2020|EPI_ISL_460317|2020- ···················· ···················· ····················· ···················· ···················· ····················
C 2732 hCoV-19/USA/MA-MGH-00015/2020|EPI_ISL_460319|2020- ···················· ···················· ····················· ···················· ···················· ····················
C 2733 hCoV-19/USA/MA-MGH-00654/2020|EPI_ISL_460330|2020- ···················· ···················· ····················· ···················· ···················· ····················
C 2734 hCoV-19/USA/MA-MGH-00148/2020|EPI_ISL_460331|2020- ···················· ···················· ····················· ···················· ···················· ····················
C 2735 hCoV-19/USA/MA-MGH-00263/2020|EPI_ISL_460332|2020- ···················· ···················· ····················· ···················· ···················· ····················
C 2736 hCoV-19/USA/MA-MGH-00428/2020|EPI_ISL_460333|2020- ···················· ···················· ····················· ···················· ···················· ····················
C 2737 hCoV-19/USA/MA-MGH-00039/2020|EPI_ISL_460334|2020- ···················· ···················· ····················· ···················· ···················· ····················
C 2738 hCoV-19/USA/MA-MGH-00056/2020|EPI_ISL_460335|2020- ···················· ···················· ····················· ···················· ···················· ····················
C 2739 hCoV-19/USA/MA-MGH-00698/2020|EPI_ISL_460336|2020- ···················· ···················· ····················· ···················· ···················· ····················
C 2740 hCoV-19/USA/MA-MGH-00582/2020|EPI_ISL_460337|2020- ···················· ···················· ····················· ···················· ···················· ····················
C 2741 hCoV-19/USA/MA-MGH-00551/2020|EPI_ISL_460338|2020- ···················· ···················· ····················· ···················· ···················· ····················
C 2742 hCoV-19/USA/MA-MGH-00155/2020|EPI_ISL_460339|2020- ···················· ···················· ····················· ···················· ···················· ····················
C 2743 hCoV-19/USA/MA-MGH-00615/2020|EPI_ISL_460340|2020- ···················· ···················· ····················· ···················· ···················· ····················
C 2744 hCoV-19/USA/MA-MGH-00673/2020|EPI_ISL_460341|2020- ···················· ···················· ····················· ···················· ···················· ····················
C 2745 hCoV-19/USA/MA-MGH-00296/2020|EPI_ISL_460342|2020- ···················· ···················· ····················· ···················· ···················· ····················
C 2746 hCoV-19/USA/MA-MGH-00492/2020|EPI_ISL_460343|2020- ···················· ···················· ····················· ···················· ···················· ····················
C 2747 hCoV-19/USA/MA-MGH-00528/2020|EPI_ISL_460344|2020- ···················· ···················· ····················· ···················· ···················· ····················
C 2748 hCoV-19/USA/MA-MGH-00231/2020|EPI_ISL_460345|2020- ···················· ···················· ····················· ···················· ···················· ····················
C 2749 hCoV-19/USA/MA-MGH-00243/2020|EPI_ISL_460346|2020- ···················· ···················· ····················· ···················· ···················· ····················
C 2750 hCoV-19/USA/MA-MGH-00516/2020|EPI_ISL_460347|2020- ···················· ···················· ····················· ···················· ···················· ····················
C 2752 hCoV-19/USA/MA-MGH-00553/2020|EPI_ISL_460349|2020- ···················· ···················· ····················· ···················· ···················· ····················
C 2753 hCoV-19/USA/MA-MGH-00318/2020|EPI_ISL_460351|2020- ···················· ···················· ····················· ···················· ···················· ····················
C 2754 hCoV-19/USA/MA-MGH-00446/2020|EPI_ISL_460352|2020- ···················· ···················· ····················· ···················· ···················· ····················
C 2755 hCoV-19/USA/MA-MGH-00416/2020|EPI_ISL_460353|2020- ···················· ···················· ····················· ···················· ···················· ····················
C 2756 hCoV-19/USA/MA-MGH-00444/2020|EPI_ISL_460354|2020- ···················· ···················· ····················· ···················· ···················· ····················
C 2757 hCoV-19/USA/MA-MGH-00670/2020|EPI_ISL_460356|2020- ···················· ···················· ····················· ···················· ···················· ····················
C 2758 hCoV-19/USA/MA-MGH-00703/2020|EPI_ISL_460357|2020- ···················· ···················· ····················· ···················· ···················· ····················
C 2759 hCoV-19/USA/MA-MGH-00701/2020|EPI_ISL_460358|2020- ···················· ···················· ····················· ···················· ···················· ····················
C 2760 hCoV-19/USA/MA-MGH-00140/2020|EPI_ISL_460359|2020- ···················· ···················· ····················· ···················· ···················· ····················
C 2761 hCoV-19/USA/MA-MGH-00261/2020|EPI_ISL_460360|2020- ···················· ···················· ····················· ···················· ···················· ····················
C 2762 hCoV-19/USA/MA-MGH-00472/2020|EPI_ISL_460361|2020- ···················· ···················· ····················· ···················· ···················· ····················
C 2763 hCoV-19/USA/MA-MGH-00183/2020|EPI_ISL_460362|2020- ···················· ···················· ····················· ···················· ···················· ····················
C 2764 hCoV-19/USA/MA-MGH-00136/2020|EPI_ISL_460364|2020- ···················· ···················· ····················· ···················· ···················· ····················
C 2765 hCoV-19/USA/MA-MGH-00238/2020|EPI_ISL_460365|2020- ···················· ···················· ····················· ···················· ···················· ····················
C 2766 hCoV-19/USA/MA-MGH-00430/2020|EPI_ISL_460366|2020- ···················· ···················· ····················· ···················· ···················· ····················
C 2767 hCoV-19/USA/MA-MGH-00194/2020|EPI_ISL_460368|2020- ···················· ···················· ····················· ···················· ···················· ····················
C 2768 hCoV-19/USA/MA-MGH-00225/2020|EPI_ISL_460369|2020- ···················· ···················· ····················· ···················· ···················· ····················
C 2769 hCoV-19/USA/MA-MGH-00232/2020|EPI_ISL_460370|2020- ···················· ···················· ····················· ···················· ···················· ····················
C 2770 hCoV-19/USA/MA-MGH-00418/2020|EPI_ISL_460371|2020- ···················· ···················· ····················· ···················· ···················· ····················
C 2771 hCoV-19/USA/MA-MGH-00189/2020|EPI_ISL_460372|2020- ···················· ···················· ····················· ···················· ···················· ····················
C 2772 hCoV-19/USA/MA-MGH-00251/2020|EPI_ISL_460373|2020- ···················· ···················· ····················· ···················· ···················· ····················
C 2773 hCoV-19/Scotland/CVR3011/2020|EPI_ISL_461620|2020- ···················· ···················· ····················· ···················· ···················· ····················
C 2774 hCoV-19/Scotland/CVR3012/2020|EPI_ISL_461621|2020- ···················· ···················· ····················· ···················· ···················· ····················
C 2775 hCoV-19/Scotland/CVR3013/2020|EPI_ISL_461622|2020- ···················· ···················· ····················· ···················· ···················· ····················
C 2776 hCoV-19/Scotland/CVR3015/2020|EPI_ISL_461623|2020- ···················· ···················· ····················· ···················· ···················· ····················
C 2779 hCoV-19/Scotland/CVR3033/2020|EPI_ISL_461626|2020- ···················· ···················· ····················· ···················· ···················· ····················
C 2780 hCoV-19/Scotland/CVR3036/2020|EPI_ISL_461628|2020- ···················· ···················A ····················· ···················· ···················· ···················· 1
C 2781 hCoV-19/Scotland/CVR3041/2020|EPI_ISL_461629|2020- ···················· ···················A ····················· ···················· ···················· ···················· 1
C 2783 hCoV-19/Scotland/CVR3043/2020|EPI_ISL_461631|2020- ···················· ···················· ····················· ···················· ···················· ····················
C 2785 hCoV-19/Scotland/CVR3047/2020|EPI_ISL_461635|2020- ···················· ···················· ····················· ···················· ···················· ····················
C 2786 hCoV-19/Scotland/CVR3049/2020|EPI_ISL_461637|2020- ···················· ···················· ····················· ···················· ···················· ····················
C 2787 hCoV-19/Scotland/CVR3050/2020|EPI_ISL_461638|2020- ···················· ···················· ····················· ···················· ···················· ····················
C 2788 hCoV-19/Scotland/CVR3051/2020|EPI_ISL_461639|2020- ···················· ··············C····· ····················· ···················· ···················· ····················
C 2789 hCoV-19/Scotland/CVR3054/2020|EPI_ISL_461641|2020- ···················· ···················· ····················· ···················· ···················· ····················
C 2790 hCoV-19/Scotland/CVR3056/2020|EPI_ISL_461642|2020- ···················· ···················· ····················· ···················· ···················· ····················

```
C    2791 hCoV-19/Scotland/CVR3057/2020|EPI_ISL_461643|2020-   ····················  ····················  ····················  ····················  ····················  ····················
C    2792 hCoV-19/Scotland/CVR3059/2020|EPI_ISL_461644|2020-   ····················  ····················  ····················  ····················  ····················  ····················
C    2793 hCoV-19/Scotland/CVR3063/2020|EPI_ISL_461647|2020-   ····················  ····················  ····················  ····················  ····················  ····················
C    2794 hCoV-19/Scotland/CVR3065/2020|EPI_ISL_461649|2020-   ····················  ····················  ····················  ····················  ····················  ····················
C    2795 hCoV-19/Scotland/CVR3077/2020|EPI_ISL_461661|2020-   ····················  ····················  ····················  ····················  ····················  ····················
C    2797 hCoV-19/Scotland/CVR3080/2020|EPI_ISL_461664|2020-   ····················  ····················  ····················  ····················  ····················  ····················
C    2798 hCoV-19/Scotland/CVR3081/2020|EPI_ISL_461665|2020-   ····················  ····················  ····················  ····················  ····················  ····················
C    2800 hCoV-19/Scotland/CVR3092/2020|EPI_ISL_461668|2020-   ····················  ····················  ····················  ····················  ····················  ····················
C    2801 hCoV-19/Scotland/CVR3096/2020|EPI_ISL_461669|2020-   ····················  ····················  ····················  ····················  ····················  ····················
C    2802 hCoV-19/Scotland/CVR3097/2020|EPI_ISL_461670|2020-   ····················  ·A··················  ····················  ····················  ····················  ····················
C    2804 hCoV-19/Scotland/CVR3102/2020|EPI_ISL_461675|2020-   ····················  ····················  ····················  ····················  ····················  ····················
C    2805 hCoV-19/Scotland/CVR3103/2020|EPI_ISL_461676|2020-   ····················  ···················A  ····················  ····················  ····················  ····················  1
C    2806 hCoV-19/Scotland/CVR3105/2020|EPI_ISL_461678|2020-   ····················  ····················  ····················  ····················  ····················  ····················
C    2807 hCoV-19/Scotland/CVR3113/2020|EPI_ISL_461685|2020-   ····················  ····················  ····················  ····················  ····················  ····················
C    2809 hCoV-19/Scotland/CVR3119/2020|EPI_ISL_461691|2020-   ····················  ····················  ····················  ····················  ····················  ····················
C    2810 hCoV-19/Scotland/CVR3136/2020|EPI_ISL_461698|2020-   ····················  ····················  ····················  ····················  ····················  ····················
C    2811 hCoV-19/Scotland/CVR3137/2020|EPI_ISL_461699|2020-   ····················  ·A··················  ····················  ····················  ····················  ····················
C    2812 hCoV-19/Spain/AS-MG-67-20/2020|EPI_ISL_462995|2020   ····················  ····················  ····················  ····················  ····················  ····················
C    2813 hCoV-19/Spain/AS-MG-68-19/2020|EPI_ISL_462996|2020   ····················  ····················  ····················  ····················  ····················  ····················
C    2814 hCoV-19/Spain/AS-MG-80-19/2020|EPI_ISL_462997|2020   ····················  ····················  ····················  ····················  ····················  ····················
C    2815 hCoV-19/Spain/AS-MG-86-17/2020|EPI_ISL_462998|2020   ····················  ····················  ····················  ····················  ····················  ····················
C    2816 hCoV-19/Spain/AS-MG-96-19/2020|EPI_ISL_462999|2020   ····················  ····················  ····················  ····················  ····················  ····················
C    2817 hCoV-19/Israel/701002442/2020|EPI_ISL_447300|2020-   ····················  ····················  ····················  ····················  ····················  ····················
C    2818 hCoV-19/Israel/701002440/2020|EPI_ISL_447301|2020-   ····················  ····················  ····················  ····················  ····················  ····················
C    2819 hCoV-19/Israel/701002431/2020|EPI_ISL_447302|2020-   ····················  ····················  ····················  ····················  ····················  ····················
C    2820 hCoV-19/Israel/701002426/2020|EPI_ISL_447303|2020-   ····················  ····················  ····················  ····················  ····················  ····················
C    2821 hCoV-19/Israel/701002403/2020|EPI_ISL_447305|2020-   ····················  ····················  ····················  ····················  ····················  ····················
C    2822 hCoV-19/Israel/701002334/2020|EPI_ISL_447306|2020-   ····················  ····················  ····················  ····················  ····················  ····················
C    2823 hCoV-19/Israel/701002327/2020|EPI_ISL_447307|2020-   ····················  ····················  ····················  ····················  ····················  ····················
C    2824 hCoV-19/Israel/701002317/2020|EPI_ISL_447308|2020-   ····················  ····················  ····················  ····················  ····················  ····················
C    2825 hCoV-19/Israel/701002314/2020|EPI_ISL_447309|2020-   ····················  ····················  ····················  ····················  ····················  ····················
C    2826 hCoV-19/Israel/701002313/2020|EPI_ISL_447310|2020-   ····················  ····················  ····················  ····················  ····················  ····················
C    2827 hCoV-19/Israel/51146669/2020|EPI_ISL_447312|2020-0   ····················  ····················  ····················  ····················  ····················  ····················
C    2828 hCoV-19/Israel/51146503/2020|EPI_ISL_447313|2020-0   ····················  ····················  ····················  ····················  ····················  ····················
C    2829 hCoV-19/Israel/51146500/2020|EPI_ISL_447314|2020-0   ····················  ····················  ····················  ····················  ····················  ····················
C    2830 hCoV-19/Israel/51146355/2020|EPI_ISL_447315|2020-0   ····················  ····················  ····················  ····················  ···N················  ····················
C    2831 hCoV-19/Israel/51145482/2020|EPI_ISL_447316|2020-0   ····················  ····················  ····················  ····················  ···A················  ····················
C    2832 hCoV-19/Israel/51145198/2020|EPI_ISL_447317|2020-0   ····················  ····················  ····················  ····················  ····················  ····················
C    2833 hCoV-19/Israel/51141225/2020|EPI_ISL_447319|2020-0   ····················  ····················  ····················  ····················  ····················  ····················
C    2834 hCoV-19/Israel/51141121/2020|EPI_ISL_447320|2020-0   ····················  ····················  ····················  ····················  ···A················  ····················
C    2835 hCoV-19/Israel/51141014/2020|EPI_ISL_447321|2020-0   ····················  ····················  ····················  ····················  ····················  ····················
C    2836 hCoV-19/Israel/51140539/2020|EPI_ISL_447323|2020-0   ····················  ····················  ····················  ····················  ····················  ····················
C    2837 hCoV-19/Israel/51140315/2020|EPI_ISL_447324|2020-0   ····················  ····················  ····················  ····················  ····················  ····················
C    2838 hCoV-19/Israel/51140068/2020|EPI_ISL_447327|2020-0   ····················  ····················  ····················  ····················  ····················  ····················
C    2839 hCoV-19/England/OXON-AD937/2020|EPI_ISL_448604|202   ····················  ····················  ····················  ····················  ····················  ····················
C    2840 hCoV-19/England/OXON-AD973/2020|EPI_ISL_448605|202   ····················  ····················  ····················  ····················  ····················  ····················
C    2841 hCoV-19/England/OXON-AD991/2020|EPI_ISL_448606|202   ····················  ····················  ····················  ····················  ····················  ····················
C    2842 hCoV-19/England/OXON-AD9BF/2020|EPI_ISL_448607|202   ····················  ····················  ····················  ····················  ····················  ····················
C    2843 hCoV-19/England/OXON-AD9DD/2020|EPI_ISL_448608|202   ····················  ····················  ····················  ····················  ····················  ····················
C    2844 hCoV-19/England/OXON-ADA16/2020|EPI_ISL_448609|202   ····················  ····················  ····················  ····················  ····················  ····················
C    2845 hCoV-19/England/OXON-ADA34/2020|EPI_ISL_448610|202   ····················  ····················  ····················  ····················  ····················  ····················
C    2846 hCoV-19/England/OXON-ADA9E/2020|EPI_ISL_448611|202   ····················  ····················  ····················  ····················  ····················  ····················
C    2847 hCoV-19/England/OXON-ADABC/2020|EPI_ISL_448612|202   ····················  ····················  ····················  ····················  ····················  ····················
C    2848 hCoV-19/England/OXON-ADADA/2020|EPI_ISL_448613|202   ····················  ····················  ····················  ····················  ····················  ····················
C    2849 hCoV-19/England/OXON-ADAF8/2020|EPI_ISL_448614|202   ····················  ····················  ····················  ····················  ····················  ····················
C    2850 hCoV-19/England/OXON-ADB31/2020|EPI_ISL_448616|202   ····················  ····················  ····················  ····················  ····················  ····················
C    2851 hCoV-19/England/OXON-ADB5F/2020|EPI_ISL_448617|202   ····················  ····················  ····················  ····················  ····················  ····················
C    2852 hCoV-19/England/OXON-ADB7D/2020|EPI_ISL_448618|202   ····················  ····················  ····················  ····················  ····················  ····················
C    2853 hCoV-19/England/OXON-ADB9B/2020|EPI_ISL_448619|202   ····················  ····················  ····················  ····················  ····················  ····················
C    2854 hCoV-19/England/OXON-ADBB9/2020|EPI_ISL_448620|202   ····················  ····················  ····················  ····················  ····················  ····················
C    2855 hCoV-19/England/OXON-ADBD7/2020|EPI_ISL_448621|202   ····················  ····················  ····················  ····················  ····················  ····················
C    2856 hCoV-19/England/OXON-ADBF5/2020|EPI_ISL_448622|202   ····················  ····················  ····················  ····················  ····················  ····················
C    2857 hCoV-19/England/OXON-ADC10/2020|EPI_ISL_448623|202   ····················  ····················  ····················  ····················  ····················  ····················
C    2858 hCoV-19/England/OXON-ADC3E/2020|EPI_ISL_448624|202   ····················  ····················  ····················  ····················  ····················  ····················
C    2859 hCoV-19/England/OXON-ADC5C/2020|EPI_ISL_448625|202   ····················  ····················  ····················  ····················  ····················  ····················
C    2860 hCoV-19/England/OXON-ADC7A/2020|EPI_ISL_448626|202   ····················  ····················  ····················  ····················  ····················  ····················
C    2861 hCoV-19/England/OXON-ADC98/2020|EPI_ISL_448627|202   ····················  ····················  ····················  ····················  ····················  ····················
C    2862 hCoV-19/England/OXON-ADCB6/2020|EPI_ISL_448628|202   ····················  ····················  ····················  ····················  ····················  ····················
C    2863 hCoV-19/England/OXON-ADCD4/2020|EPI_ISL_448629|202   ····················  ····················  ····················  ····················  ····················  ····················
C    2864 hCoV-19/England/OXON-ADCF2/2020|EPI_ISL_448630|202   ····················  ····················  ····················  ····················  ····················  ····················
C    2865 hCoV-19/England/OXON-ADD1D/2020|EPI_ISL_448631|202   ····················  ····················  ····················  ····················  ····················  ····················
C    2866 hCoV-19/England/OXON-ADD3B/2020|EPI_ISL_448632|202   ····················  ····················  ····················  ····················  ····················  ····················
C    2867 hCoV-19/England/OXON-ADD59/2020|EPI_ISL_448633|202   ····················  ····················  ····················  ····················  ····················  ····················
C    2868 hCoV-19/England/OXON-ADD77/2020|EPI_ISL_448634|202   ····················  ····················  ····················  ····················  ····················  ····················
C    2869 hCoV-19/England/OXON-ADD95/2020|EPI_ISL_448635|202   ····················  ····················  ····················  ····················  ····················  ····················
C    2870 hCoV-19/England/OXON-ADDB3/2020|EPI_ISL_448636|202   ····················  ····················  ····················  ····················  ····················  ····················
C    2871 hCoV-19/England/OXON-ADDD1/2020|EPI_ISL_448637|202   ····················  ····················  ····················  ····················  ····················  ····················
C    2872 hCoV-19/England/OXON-ADDFF/2020|EPI_ISL_448638|202   ····················  ····················  ····················  ····················  ····················  ····················
```

| | | | | | | | | |
|---|---|---|---|---|---|---|---|---|
| C | 2873 | hCoV-19/England/OXON-ADE1A/2020\|EPI_ISL_448639\|202 | .................... | .................... | .................... | .................... | .................... | .................... |
| C | 2874 | hCoV-19/England/OXON-ADE38/2020\|EPI_ISL_448640\|202 | .................... | .................... | .................... | .................... | .................... | .................... |
| C | 2875 | hCoV-19/England/OXON-ADEB0/2020\|EPI_ISL_448643\|202 | .................... | .................... | .................... | .................... | .................... | .................... |
| C | 2876 | hCoV-19/England/OXON-ADEDE/2020\|EPI_ISL_448644\|202 | .................... | .................... | .................... | .................... | .................... | .................... |
| C | 2877 | hCoV-19/England/OXON-ADEFC/2020\|EPI_ISL_448645\|202 | .................... | .................... | .................... | .................... | .................... | .................... |
| C | 2878 | hCoV-19/England/OXON-ADF17/2020\|EPI_ISL_448646\|202 | .................... | .................... | .................... | .................... | .................... | .................... |
| C | 2879 | hCoV-19/England/OXON-ADF71/2020\|EPI_ISL_448650\|202 | .................... | .................... | .................... | .................... | .................... | .................... |
| C | 2880 | hCoV-19/England/OXON-ADF9F/2020\|EPI_ISL_448651\|202 | .................... | .................... | .................... | .................... | .................... | .................... |
| C | 2881 | hCoV-19/England/OXON-ADFBD/2020\|EPI_ISL_448652\|202 | .................... | .................... | .................... | .................... | .................... | .................... |
| C | 2882 | hCoV-19/England/OXON-ADFF9/2020\|EPI_ISL_448654\|202 | .................... | .................... | .................... | .................... | .................... | .................... |
| C | 2883 | hCoV-19/England/OXON-AE013/2020\|EPI_ISL_448655\|202 | .................... | .................... | .................... | .................... | .................... | .................... |
| C | 2884 | hCoV-19/England/OXON-AE022/2020\|EPI_ISL_448656\|202 | .................... | .................... | .................... | .................... | .................... | .................... |
| C | 2885 | hCoV-19/England/OXON-AE031/2020\|EPI_ISL_448657\|202 | .................... | .................... | .................... | .................... | .................... | .................... |
| C | 2886 | hCoV-19/England/OXON-AE05F/2020\|EPI_ISL_448658\|202 | .................... | .................... | .................... | .................... | .................... | .................... |
| C | 2887 | hCoV-19/USA/WA-S732/2020\|EPI_ISL_449902\|2020-04-13 | .................... | .................... | .................... | .................... | .................... | .................... |
| C | 2888 | hCoV-19/USA/WA-S733/2020\|EPI_ISL_449903\|2020-04-13 | .................... | .................... | .................... | .................... | .................... | .................... |
| C | 2889 | hCoV-19/USA/WA-S734/2020\|EPI_ISL_449904\|2020-04-13 | .................... | .................... | .................... | .................... | .................... | .................... |
| C | 2890 | hCoV-19/USA/WA-S735/2020\|EPI_ISL_449905\|2020-04-13 | .................... | .................... | .................... | .................... | .................... | .................... |
| C | 2891 | hCoV-19/USA/WA-S736/2020\|EPI_ISL_449906\|2020-04-13 | .................... | .................... | .................... | .................... | .................... | .................... |
| C | 2892 | hCoV-19/USA/WA-S737/2020\|EPI_ISL_449907\|2020-04-13 | .................... | .................... | .................... | .................... | .................... | .................... |
| C | 2893 | hCoV-19/USA/WA-S738/2020\|EPI_ISL_449908\|2020-04-13 | .................... | .................... | .................... | .................... | .................... | .................... |
| C | 2894 | hCoV-19/USA/WA-S739/2020\|EPI_ISL_449909\|2020-04-13 | .................... | .................... | .................... | .................... | .................... | .................... |
| C | 2895 | hCoV-19/USA/WA-S740/2020\|EPI_ISL_449910\|2020-04-13 | .................... | .................... | .................... | .................... | .................... | .................... |
| C | 2896 | hCoV-19/USA/WA-S741/2020\|EPI_ISL_449911\|2020-04-13 | .................... | .................... | .................... | .................... | .................... | .................... |
| C | 2897 | hCoV-19/USA/WA-S742/2020\|EPI_ISL_449912\|2020-04-13 | .................... | .................... | .................... | .................... | .................... | .................... |
| C | 2898 | hCoV-19/USA/WA-S743/2020\|EPI_ISL_449913\|2020-04-13 | .................... | .................... | .................... | .................... | .................... | .................... |
| C | 2899 | hCoV-19/USA/WA-S744/2020\|EPI_ISL_449914\|2020-04-12 | .................... | .................... | .................... | .................... | .................... | .................... |
| C | 2900 | hCoV-19/USA/WA-S745/2020\|EPI_ISL_449915\|2020-04-13 | .................... | .................... | .................... | .................... | .................... | .................... |
| C | 2901 | hCoV-19/USA/WA-S747/2020\|EPI_ISL_449917\|2020-04-13 | .................... | .................... | .................... | .................... | .................... | .................... |
| C | 2902 | hCoV-19/USA/WA-S748/2020\|EPI_ISL_449918\|2020-04-13 | .................... | .................... | .................... | .................... | .................... | .................... |
| C | 2903 | hCoV-19/USA/WA-S749/2020\|EPI_ISL_449919\|2020-04-12 | .................... | .................... | .................... | .................... | .................... | .................... |
| C | 2904 | hCoV-19/USA/WA-S750/2020\|EPI_ISL_449920\|2020-04-13 | .................... | .................... | .................... | .................... | .................... | .................... |
| C | 2905 | hCoV-19/USA/WA-S751/2020\|EPI_ISL_449921\|2020-04-11 | .................... | .................... | .................... | .................... | .................... | .................... |
| C | 2906 | hCoV-19/USA/WA-S752/2020\|EPI_ISL_449922\|2020-04-13 | .................... | .................... | .................... | .................... | .................... | .................... |
| C | 2907 | hCoV-19/USA/WA-S753/2020\|EPI_ISL_449923\|2020-04-13 | .................... | .................... | .................... | .................... | .................... | .................... |
| C | 2908 | hCoV-19/USA/WA-S756/2020\|EPI_ISL_449926\|2020-04-14 | .................... | .................... | .................... | .................... | .................... | .................... |
| C | 2909 | hCoV-19/USA/WA-S757/2020\|EPI_ISL_449927\|2020-04-13 | .................... | .................... | .................... | .................... | .................... | .................... |
| C | 2910 | hCoV-19/USA/WA-S765/2020\|EPI_ISL_449935\|2020-04-14 | .................... | .................... | .................... | .................... | .................... | .................... |
| C | 2911 | hCoV-19/USA/WA-S766/2020\|EPI_ISL_449936\|2020-04-14 | .................... | .................... | .................... | .................... | .................... | .................... |
| C | 2912 | hCoV-19/USA/WA-S767/2020\|EPI_ISL_449937\|2020-04-14 | .................... | .................... | .................... | .................... | .................... | .................... |
| C | 2913 | hCoV-19/USA/WA-S768/2020\|EPI_ISL_449938\|2020-04-14 | .................... | .................... | .................... | .................... | .................... | .................... |
| C | 2914 | hCoV-19/USA/WA-S769/2020\|EPI_ISL_449939\|2020-04-14 | .................... | .................... | .................... | .................... | .................... | .................... |
| C | 2915 | hCoV-19/USA/WA-S770/2020\|EPI_ISL_449940\|2020-04-13 | .................... | .................... | .................... | .................... | .................... | .................... |
| C | 2916 | hCoV-19/USA/WA-S771/2020\|EPI_ISL_449941\|2020-04-13 | .................... | .................... | .................... | .................... | .................... | .................... |
| C | 2917 | hCoV-19/USA/WA-S773/2020\|EPI_ISL_449943\|2020-04-14 | .................... | .................... | .................... | .................... | .................... | .................... |
| C | 2918 | hCoV-19/USA/WA-S774/2020\|EPI_ISL_449944\|2020-04-14 | .................... | .................... | .................... | .................... | .................... | .................... |
| C | 2919 | hCoV-19/USA/WA-S775/2020\|EPI_ISL_449945\|2020-04-14 | .................... | .................... | .................... | .................... | .................... | .................... |
| C | 2920 | hCoV-19/USA/WA-S776/2020\|EPI_ISL_449946\|2020-04-14 | .................... | .................... | .................... | .................... | .................... | .................... |
| C | 2921 | hCoV-19/USA/WA-S777/2020\|EPI_ISL_449947\|2020-04-14 | .................... | .................... | .................... | .................... | .................... | .................... |
| C | 2922 | hCoV-19/USA/WA-S778/2020\|EPI_ISL_449948\|2020-04-14 | .................... | .................... | .................... | .................... | .................... | .................... |
| C | 2923 | hCoV-19/USA/WA-S779/2020\|EPI_ISL_449949\|2020-04-14 | .................... | .................... | .................... | .................... | .................... | .................... |
| C | 2924 | hCoV-19/USA/WA-S781/2020\|EPI_ISL_449951\|2020-04-14 | .................... | .................... | .................... | .................... | .................... | .................... |
| C | 2925 | hCoV-19/USA/WA-S782/2020\|EPI_ISL_449952\|2020-04-14 | .................... | .................... | .................... | .................... | .................... | .................... |
| C | 2926 | hCoV-19/USA/WA-S784/2020\|EPI_ISL_449954\|2020-04-14 | .................... | .................... | .................... | .................... | .................... | .................... |
| C | 2927 | hCoV-19/USA/WA-S785/2020\|EPI_ISL_449955\|2020-04-14 | .................... | .................... | .................... | .................... | .................... | .................... |
| C | 2928 | hCoV-19/USA/WA-S786/2020\|EPI_ISL_449956\|2020-04-14 | .................... | .................... | .................... | .................... | .................... | .................... |
| C | 2929 | hCoV-19/USA/WA-S787/2020\|EPI_ISL_449957\|2020-04-14 | .................... | .................... | .................... | .................... | .................... | .................... |
| C | 2930 | hCoV-19/USA/WA-S788/2020\|EPI_ISL_449958\|2020-04-14 | .................... | .................... | .................... | .................... | .................... | .................... |
| C | 2931 | hCoV-19/USA/WA-S789/2020\|EPI_ISL_449959\|2020-04-15 | .................... | .................... | .................... | .................... | .................... | .................... |
| C | 2932 | hCoV-19/USA/WA-S790/2020\|EPI_ISL_449960\|2020-04-15 | .................... | .................... | .................... | .................... | .................... | .................... |
| C | 2933 | hCoV-19/USA/WA-S792/2020\|EPI_ISL_449962\|2020-04-15 | .................... | .................... | .................... | .................... | .................... | .................... |
| C | 2934 | hCoV-19/USA/WA-S793/2020\|EPI_ISL_449963\|2020-04-15 | .................... | .................... | .................... | .................... | .................... | .................... |
| C | 2935 | hCoV-19/USA/WA-S794/2020\|EPI_ISL_449964\|2020-04-14 | .................... | .................... | .................... | .................... | .................... | .................... |
| C | 2936 | hCoV-19/USA/WA-S795/2020\|EPI_ISL_449965\|2020-04-14 | .................... | .................... | .................... | .................... | .................... | .................... |
| C | 2937 | hCoV-19/USA/WA-S796/2020\|EPI_ISL_449966\|2020-04-15 | .................... | .................... | .................... | .................... | .................... | .................... |
| C | 2938 | hCoV-19/USA/WA-S797/2020\|EPI_ISL_449967\|2020-04-16 | .................... | .................... | .................... | .................... | .................... | .................... |
| C | 2939 | hCoV-19/USA/WA-S798/2020\|EPI_ISL_449968\|2020-04-16 | .................... | .................... | .................... | .................... | .................... | .................... |
| C | 2940 | hCoV-19/USA/WA-S799/2020\|EPI_ISL_449969\|2020-04-16 | .................... | .................... | .................... | .................... | .................... | .................... |
| C | 2941 | hCoV-19/USA/WA-S800/2020\|EPI_ISL_449970\|2020-04-16 | .................... | .................... | .................... | .................... | .................... | .................... |
| C | 2942 | hCoV-19/USA/WA-S801/2020\|EPI_ISL_449971\|2020-04-16 | .................... | .................... | .................... | .................... | .................... | .................... |
| C | 2943 | hCoV-19/USA/WA-S802/2020\|EPI_ISL_449972\|2020-04-16 | .................... | .................... | .................... | .................... | .................... | .................... |
| C | 2944 | hCoV-19/USA/WA-S803/2020\|EPI_ISL_449973\|2020-04-16 | .................... | .................... | .................... | .................... | .................... | .................... |
| C | 2945 | hCoV-19/USA/WA-S804/2020\|EPI_ISL_449974\|2020-04-15 | .................... | .................... | .................... | .................... | .................... | .................... |
| C | 2946 | hCoV-19/USA/WA-S805/2020\|EPI_ISL_449975\|2020-04-15 | .................... | .................... | .................... | .................... | .................... | .................... |
| C | 2947 | hCoV-19/USA/WA-S807/2020\|EPI_ISL_449977\|2020-04-15 | .................... | .................... | .................... | .................... | .................... | .................... |
| C | 2948 | hCoV-19/USA/WA-S808/2020\|EPI_ISL_449978\|2020-04-15 | .................... | .................... | .................... | .................... | .................... | .................... |
| C | 2949 | hCoV-19/USA/WA-S809/2020\|EPI_ISL_449979\|2020-04-15 | .................... | .................... | .................... | .................... | .................... | .................... |
| C | 2950 | hCoV-19/USA/WA-S810/2020\|EPI_ISL_449980\|2020-04-15 | .................... | .................... | .................... | .................... | .................... | .................... |

| | | | | | | | | |
|---|---|---|---|---|---|---|---|---|
| C | 2951 hCoV-19/USA/WA-S811/2020\|EPI_ISL_449981\|2020-04-16 | .................... | .................... | .................... | .................... | .................... | .................... | |
| C | 2952 hCoV-19/USA/WA-S812/2020\|EPI_ISL_449982\|2020-04-15 | .................... | .................... | .................... | .................... | .................... | .................... | |
| C | 2953 hCoV-19/USA/WA-S813/2020\|EPI_ISL_449983\|2020-04-15 | .................... | .................... | .................... | .................... | .................... | .................... | |
| C | 2954 hCoV-19/USA/WA-S814/2020\|EPI_ISL_449984\|2020-04-17 | .................... | .................... | .................... | .................... | .................... | .................... | |
| C | 2955 hCoV-19/USA/WA-S815/2020\|EPI_ISL_449985\|2020-04-17 | .................... | .................... | .................... | .................... | .................... | .................... | |
| C | 2956 hCoV-19/USA/WA-S816/2020\|EPI_ISL_449986\|2020-04-17 | .................... | .................... | .................... | .................... | .................... | .................... | |
| C | 2957 hCoV-19/USA/WA-S817/2020\|EPI_ISL_449987\|2020-04-17 | .................... | .................... | .................... | .................... | .................... | .................... | |
| C | 2958 hCoV-19/USA/WA-S818/2020\|EPI_ISL_449988\|2020-04-17 | .................... | .................... | .................... | .................... | .................... | .................... | |
| C | 2959 hCoV-19/USA/WA-S819/2020\|EPI_ISL_449989\|2020-04-15 | .................... | .................... | .................... | .................... | .................... | .................... | |
| C | 2960 hCoV-19/Denmark/ALAB-HH-122/2020\|EPI_ISL_452003\|20 | .................... | .................... | .................... | .................... | .................... | .................... | |
| C | 2961 hCoV-19/Denmark/ALAB-HH-123/2020\|EPI_ISL_452004\|20 | .................... | .................... | .................... | .................... | .................... | .................... | |
| C | 2962 hCoV-19/Denmark/ALAB-HH-124/2020\|EPI_ISL_452005\|20 | .................... | .................... | .................... | .................... | .................... | .................... | |
| C | 2963 hCoV-19/Denmark/ALAB-HH-126/2020\|EPI_ISL_452007\|20 | .................... | .................... | .................... | .................... | .................... | .................... | |
| C | 2964 hCoV-19/Denmark/ALAB-HH-127/2020\|EPI_ISL_452008\|20 | .................... | .................... | .................... | .................... | .................... | .................... | |
| C | 2965 hCoV-19/Denmark/ALAB-HH-128/2020\|EPI_ISL_452009\|20 | .................... | .................... | .................... | .................... | .................... | .................... | |
| C | 2966 hCoV-19/Denmark/ALAB-HH-129/2020\|EPI_ISL_452010\|20 | .................... | .................... | .................... | .................... | .................... | .................... | |
| C | 2967 hCoV-19/Denmark/ALAB-HH-131/2020\|EPI_ISL_452012\|20 | .................... | .................... | .................... | .................... | .................... | .................... | |
| C | 2968 hCoV-19/Denmark/ALAB-HH-134/2020\|EPI_ISL_452013\|20 | .................... | .................... | .................... | .................... | .................... | .................... | |
| C | 2969 hCoV-19/Denmark/ALAB-HH-135/2020\|EPI_ISL_452014\|20 | .................... | .................... | .................... | .................... | .................... | .................... | |
| C | 2970 hCoV-19/Denmark/ALAB-HH-136/2020\|EPI_ISL_452015\|20 | .................... | .................... | .................... | .................... | .................... | .................... | |
| C | 2971 hCoV-19/Denmark/ALAB-HH-137/2020\|EPI_ISL_452016\|20 | .................... | .................... | .................... | .................... | .................... | .................... | |
| C | 2972 hCoV-19/Denmark/ALAB-HH-138/2020\|EPI_ISL_452017\|20 | .................... | .................... | .................... | .................... | .................... | .................... | |
| C | 2973 hCoV-19/Denmark/ALAB-HH-139/2020\|EPI_ISL_452018\|20 | .................... | .................... | .................... | .................... | .................... | .................... | |
| C | 2974 hCoV-19/Denmark/ALAB-HH-140/2020\|EPI_ISL_452019\|20 | .................... | .................... | .................... | .................... | .................... | .................... | |
| C | 2975 hCoV-19/Denmark/ALAB-HH-141/2020\|EPI_ISL_452020\|20 | .................... | .................... | .................... | .................... | .................... | .................... | |
| C | 2976 hCoV-19/Denmark/ALAB-HH-142/2020\|EPI_ISL_452021\|20 | .................... | .................... | .................... | .................... | .................... | .................... | |
| C | 2977 hCoV-19/Denmark/ALAB-HH-143/2020\|EPI_ISL_452022\|20 | .................... | .................... | .................... | .................... | .................... | .................... | |
| C | 2978 hCoV-19/Denmark/ALAB-HH-144/2020\|EPI_ISL_452023\|20 | .................... | .................... | .................... | .................... | .................... | .................... | |
| C | 2979 hCoV-19/Denmark/ALAB-HH-145/2020\|EPI_ISL_452024\|20 | .................... | .................... | .................... | .................... | .................... | .................... | |
| C | 2980 hCoV-19/Denmark/ALAB-HH-148/2020\|EPI_ISL_452025\|20 | .................... | .................... | .................... | .................... | .................... | .................... | |
| C | 2981 hCoV-19/Denmark/ALAB-HH-150/2020\|EPI_ISL_452026\|20 | .................... | .................... | .................... | .................... | .................... | .................... | |
| C | 2982 hCoV-19/Denmark/ALAB-HH-151/2020\|EPI_ISL_452027\|20 | .................... | .................... | .................... | .................... | .................... | .................... | |
| C | 2983 hCoV-19/Denmark/ALAB-HH-153/2020\|EPI_ISL_452028\|20 | .................... | .................... | .................... | .................... | .................... | .................... | |
| C | 2984 hCoV-19/Denmark/ALAB-HH-154/2020\|EPI_ISL_452029\|20 | .................... | .................... | .................... | .................... | .................... | .................... | |
| C | 2985 hCoV-19/Denmark/ALAB-HH-155/2020\|EPI_ISL_452030\|20 | .................... | .................... | .................... | .................... | .................... | .................... | |
| C | 2986 hCoV-19/Denmark/ALAB-HH-156/2020\|EPI_ISL_452031\|20 | .................... | .................... | .................... | .................... | .................... | .................... | |
| C | 2987 hCoV-19/Denmark/ALAB-HH-157/2020\|EPI_ISL_452032\|20 | .................... | .................... | .................... | .................... | .................... | .................... | |
| C | 2988 hCoV-19/Denmark/ALAB-HH-158/2020\|EPI_ISL_452033\|20 | .................... | .................... | .................... | .................... | .................... | .................... | |
| C | 2989 hCoV-19/Denmark/ALAB-HH-159/2020\|EPI_ISL_452034\|20 | .................... | .................... | .................... | .................... | .................... | .................... | |
| C | 2990 hCoV-19/Denmark/ALAB-HH-163/2020\|EPI_ISL_452035\|20 | .................... | .................... | .................... | .................... | .................... | .................... | |
| C | 2994 hCoV-19/Scotland/EDB4885/2020\|EPI_ISL_453172\|2020- | .................... | .A.................. | .................... | .................... | .................... | .................... | |
| C | 2997 hCoV-19/Scotland/EDB4993/2020\|EPI_ISL_453177\|2020- | .................... | .................... | .................... | .................... | .................... | .................... | |
| C | 2998 hCoV-19/Scotland/EDB4995/2020\|EPI_ISL_453178\|2020- | .................... | .................... | .................... | .................... | .................... | .................... | |
| C | 2999 hCoV-19/Scotland/EDB4998/2020\|EPI_ISL_453179\|2020- | .................... | .A.................. | .................... | .................... | .................... | .................... | |
| C | 3000 hCoV-19/Scotland/EDB5011/2020\|EPI_ISL_453180\|2020- | .................... | .................... | .................... | .................... | .................... | .................... | |
| C | 3002 hCoV-19/Scotland/EDB5020/2020\|EPI_ISL_453182\|2020- | .................... | .................... | .................... | .................... | .................... | .................... | |
| C | 3003 hCoV-19/Scotland/EDB5027/2020\|EPI_ISL_453183\|2020- | .................... | .A.................. | .................... | .................... | .................... | .................... | |
| C | 3004 hCoV-19/Scotland/EDB5104/2020\|EPI_ISL_453184\|2020- | .................... | .................... | .................... | .................... | .................... | .................... | |
| C | 3005 hCoV-19/Scotland/EDB5105/2020\|EPI_ISL_453185\|2020- | .................... | .................... | .................... | .................... | .................... | .................... | |
| C | 3006 hCoV-19/Scotland/EDB5107/2020\|EPI_ISL_453187\|2020- | .................... | .................... | .................... | .................... | .................... | .................... | |
| C | 3007 hCoV-19/Scotland/EDB5108/2020\|EPI_ISL_453188\|2020- | .................... | .................... | .................... | .................... | .................... | .................... | |
| C | 3008 hCoV-19/Scotland/EDB5110/2020\|EPI_ISL_453190\|2020- | .................... | .................... | .................... | .................... | .................... | .................... | |
| C | 3009 hCoV-19/Scotland/EDB5112/2020\|EPI_ISL_453191\|2020- | .................... | .................... | .................... | .................... | .................... | .................... | |
| C | 3010 hCoV-19/Scotland/EDB5113/2020\|EPI_ISL_453192\|2020- | .................... | .................... | .................... | .................... | .................... | .................... | |
| C | 3011 hCoV-19/Scotland/EDB5121/2020\|EPI_ISL_453193\|2020- | .................... | .A.................. | .................... | .................... | .................... | .................... | |
| C | 3012 hCoV-19/Scotland/EDB5133/2020\|EPI_ISL_453194\|2020- | .................... | .................... | .................... | .................... | .................... | .................... | |
| C | 3013 hCoV-19/England/LIVE-9E73F/2020\|EPI_ISL_453327\|202 | .................... | ...................A | .................... | .................... | .................... | .................... | 1 |
| C | 3014 hCoV-19/USA/CA-CZB-1234/2020\|EPI_ISL_454654\|2020-0 | .................... | .................... | .................... | .................... | .................... | .................... | |
| C | 3015 hCoV-19/USA/CA-CZB-1235/2020\|EPI_ISL_454655\|2020-0 | .................... | .................... | .................... | .................... | .................... | .................... | |
| C | 3016 hCoV-19/USA/CA-CZB-1236/2020\|EPI_ISL_454656\|2020-0 | .................... | .................... | .................... | .................... | .................... | .................... | |
| C | 3017 hCoV-19/USA/CA-CZB-1237/2020\|EPI_ISL_454657\|2020-0 | .................... | .................... | .................... | .................... | .................... | .................... | |
| C | 3018 hCoV-19/USA/CA-CZB-1238/2020\|EPI_ISL_454658\|2020-0 | .................... | .................... | .................... | .................... | .................... | .................... | |
| C | 3019 hCoV-19/USA/CA-CZB-1239/2020\|EPI_ISL_454659\|2020-0 | .................... | .................... | .................... | .................... | .................... | .................... | |
| C | 3020 hCoV-19/USA/CA-CZB-1245/2020\|EPI_ISL_454665\|2020-0 | .................... | .................... | .................... | .................... | .................... | .................... | |
| C | 3021 hCoV-19/USA/CA-CZB-1246/2020\|EPI_ISL_454666\|2020-0 | .................... | .................... | .................... | .................... | .................... | .................... | |
| C | 3022 hCoV-19/USA/CA-CZB-1247/2020\|EPI_ISL_454667\|2020-0 | .................... | .................... | .................... | .................... | .................... | .................... | |
| C | 3023 hCoV-19/USA/CA-CZB-1248/2020\|EPI_ISL_454668\|2020-0 | .................... | .................... | .................... | .................... | .................... | .................... | |
| C | 3024 hCoV-19/USA/CA-CZB-1250/2020\|EPI_ISL_454670\|2020-0 | .................... | .................... | .................... | .................... | .................... | .................... | |
| C | 3025 hCoV-19/USA/CA-CZB-1252/2020\|EPI_ISL_454672\|2020-0 | .................... | .................... | .................... | .................... | .................... | .................... | |
| C | 3026 hCoV-19/USA/CA-CZB-1253/2020\|EPI_ISL_454673\|2020-0 | .................... | .................... | .................... | .................... | .................... | .................... | |
| C | 3027 hCoV-19/USA/CA-CZB-1254/2020\|EPI_ISL_454674\|2020-0 | .................... | .................... | .................... | .................... | .................... | .................... | |
| C | 3028 hCoV-19/USA/CA-CZB-1255/2020\|EPI_ISL_454675\|2020-0 | .................... | .................... | .................... | .................... | .................... | .................... | |
| C | 3029 hCoV-19/USA/CA-CZB-1256/2020\|EPI_ISL_454676\|2020-0 | .................... | .................... | .................... | .................... | .................... | .................... | |
| C | 3030 hCoV-19/USA/CA-CZB-1257/2020\|EPI_ISL_454677\|2020-0 | .................... | .................... | .................... | .................... | .................... | .................... | |
| C | 3031 hCoV-19/USA/CA-CZB-1258/2020\|EPI_ISL_454678\|2020-0 | .................... | .................... | .................... | .................... | .................... | .................... | |
| C | 3032 hCoV-19/USA/CA-CZB-1259/2020\|EPI_ISL_454679\|2020-0 | .................... | .................... | .................... | .................... | .................... | .................... | |
| C | 3033 hCoV-19/USA/CA-CZB-1260/2020\|EPI_ISL_454680\|2020-0 | .................... | .................... | .................... | .................... | .................... | .................... | |
| C | 3034 hCoV-19/USA/CA-CZB-1261/2020\|EPI_ISL_454681\|2020-0 | .................... | .................... | .................... | .................... | .................... | .................... | |

| | | | | | | | | |
|---|---|---|---|---|---|---|---|---|
| C | 3035 hCoV-19/USA/CA-CZB-1262/2020\|EPI_ISL_454682\|2020-0 | ···················· | ···················· | ···················· | ···················· | ···················· | ···················· | |
| C | 3036 hCoV-19/USA/CA-CZB-1263/2020\|EPI_ISL_454683\|2020-0 | ···················· | ···················· | ···················· | ···················· | ···················· | ···················· | |
| C | 3037 hCoV-19/USA/CA-CZB-1264/2020\|EPI_ISL_454684\|2020-0 | ···················· | ···················· | ···················· | ···················· | ···················· | ···················· | |
| C | 3038 hCoV-19/USA/CA-CZB-1265/2020\|EPI_ISL_454685\|2020-0 | ···················· | ···················· | ···················· | ···················· | ···················· | ···················· | |
| C | 3039 hCoV-19/USA/CA-CZB-1266/2020\|EPI_ISL_454686\|2020-0 | ···················· | ···················· | ···················· | ···················· | ···················· | ···················· | |
| C | 3040 hCoV-19/USA/CA-CZB-1267/2020\|EPI_ISL_454687\|2020-0 | ···················· | ···················· | ···················· | ···················· | ···················· | ···················· | |
| C | 3041 hCoV-19/USA/CA-CZB-1268/2020\|EPI_ISL_454688\|2020-0 | ···················· | ···················· | ···················· | ···················· | ···················· | ···················· | |
| C | 3042 hCoV-19/USA/CA-CZB-1269/2020\|EPI_ISL_454689\|2020-0 | ···················· | ···················· | ···················· | ···················· | ···················· | ···················· | |
| C | 3043 hCoV-19/USA/GA-EHC-083E/2020\|EPI_ISL_454690\|2020-0 | ···················· | ···················· | ···················· | ···················· | ···················· | ···················· | |
| C | 3044 hCoV-19/USA/NY-QDX-01/2020\|EPI_ISL_454691\|2020-03- | ···················· | ···················· | ···················· | ···················· | ···················· | ···················· | |
| C | 3045 hCoV-19/USA/LA-EVTL142/2020\|EPI_ISL_455985\|2020-03 | ···················· | ···················· | ···················· | ···················· | ···················· | ···················· | |
| C | 3046 hCoV-19/USA/LA-EVTL143/2020\|EPI_ISL_455986\|2020-03 | ···················· | ···················· | ···················· | ···················· | ···················· | ···················· | |
| C | 3047 hCoV-19/USA/LA-EVTL144/2020\|EPI_ISL_455987\|2020-03 | ···················· | ···················· | ···················· | ···················· | ···················· | ···················· | |
| C | 3048 hCoV-19/USA/LA-EVTL145/2020\|EPI_ISL_455988\|2020-03 | ···················· | ···················· | ···················· | ···················· | ···················· | ···················· | |
| C | 3049 hCoV-19/USA/LA-EVTL146/2020\|EPI_ISL_455989\|2020-03 | ···················· | ···················· | ···················· | ···················· | ···················· | ···················· | |
| C | 3050 hCoV-19/USA/LA-EVTL147/2020\|EPI_ISL_455990\|2020-03 | ···················· | ···················· | ···················· | ···················· | ···················· | ···················· | |
| C | 3051 hCoV-19/USA/LA-EVTL148/2020\|EPI_ISL_455991\|2020-03 | ···················· | ···················· | ···················· | ···················· | ···················· | ···················· | |
| C | 3052 hCoV-19/USA/LA-EVTL153/2020\|EPI_ISL_455996\|2020-04 | ···················· | ···················· | ···················· | ···················· | ···················· | ···················· | |
| C | 3053 hCoV-19/USA/LA-EVTL154/2020\|EPI_ISL_455997\|2020-04 | ···················· | ···················· | ···················· | ···················· | ···················· | ···················· | |
| C | 3054 hCoV-19/USA/LA-EVTL155/2020\|EPI_ISL_455998\|2020-04 | ···················· | ···················· | ···················· | ···················· | ···················· | ···················· | |
| C | 3055 hCoV-19/USA/LA-EVTL156/2020\|EPI_ISL_455999\|2020-04 | ···················· | ···················· | ···················· | ···················· | ···················· | ···················· | |
| C | 3056 hCoV-19/Scotland/CVR2940/2020\|EPI_ISL_461600\|2020- | ···················· | ···················· | ···················· | ···················· | ···················· | ···················· | |
| C | 3057 hCoV-19/Scotland/CVR2941/2020\|EPI_ISL_461601\|2020- | ···················· | ···················· | ···················· | ···················· | ···················· | ···················· | |
| C | 3058 hCoV-19/Scotland/CVR2947/2020\|EPI_ISL_461603\|2020- | ···················· | ···················· | ···················· | ···················· | ···················· | ···················· | |
| C | 3059 hCoV-19/Scotland/CVR2948/2020\|EPI_ISL_461604\|2020- | ···················· | ··············C····· | ···················· | ···················· | ···················· | ···················· | |
| C | 3060 hCoV-19/Scotland/CVR2950/2020\|EPI_ISL_461605\|2020- | ···················· | ···················A | ···················· | ···················· | ···················· | ···················· | 1 |
| C | 3061 hCoV-19/Scotland/CVR2951/2020\|EPI_ISL_461606\|2020- | ···················· | ···················· | ···················· | ···················· | ···················· | ···················· | |
| C | 3062 hCoV-19/Scotland/CVR2953/2020\|EPI_ISL_461607\|2020- | ···················· | ···················· | ···················· | ···················· | ···················· | ···················· | |
| C | 3063 hCoV-19/Scotland/CVR2957/2020\|EPI_ISL_461610\|2020- | ···················· | ···················· | ···················· | ···················· | ···················· | ···················· | |
| C | 3064 hCoV-19/Scotland/CVR2966/2020\|EPI_ISL_461611\|2020- | ···················· | ···················· | ···················· | ···················· | ···················· | ···················· | |
| C | 3065 hCoV-19/Scotland/CVR2991/2020\|EPI_ISL_461612\|2020- | ···················· | ···················A | ···················· | ···················· | ···················· | ···················· | 1 |
| C | 3066 hCoV-19/Scotland/CVR2996/2020\|EPI_ISL_461613\|2020- | ···················· | ···················A | ···················· | ···················· | ···················· | ···················· | 1 |
| C | 3067 hCoV-19/Scotland/CVR2997/2020\|EPI_ISL_461614\|2020- | ···················· | ·A·················· | ···················· | ···················· | ···················· | ···················· | |
| C | 3068 hCoV-19/Scotland/CVR2999/2020\|EPI_ISL_461615\|2020- | ···················· | ···················· | ···················· | ···················· | ···················· | ···················· | |
| C | 3069 hCoV-19/Scotland/CVR3002/2020\|EPI_ISL_461616\|2020- | ···················· | ···················· | ···················· | ···················· | ···················· | ···················· | |
| C | 3071 hCoV-19/Scotland/CVR3009/2020\|EPI_ISL_461618\|2020- | ···················· | ···················· | ···················· | ···················· | ···················· | ···················· | |
| C | 3072 hCoV-19/Scotland/CVR3010/2020\|EPI_ISL_461619\|2020- | ···················· | ···················· | ···················· | ···················· | ···················· | ···················· | |
| C | 3073 hCoV-19/Israel/130711367/2020\|EPI_ISL_447452\|2020- | ···················· | ···················· | ···················· | ···················· | ···················· | ···················· | |
| C | 3074 hCoV-19/Israel/130711116/2020\|EPI_ISL_447453\|2020- | ···················· | ···················· | ···················· | ···················· | ···················· | ···················· | |
| C | 3075 hCoV-19/Israel/130711112/2020\|EPI_ISL_447454\|2020- | ···················· | ···················· | ···················· | ···················· | ···N················ | ···················· | |
| C | 3077 hCoV-19/Israel/130711082/2020\|EPI_ISL_447456\|2020- | ···················· | ···················· | ···················· | ···················· | ···················· | ···················· | |
| C | 3078 hCoV-19/Israel/130710716/2020\|EPI_ISL_447457\|2020- | ···················· | ···················· | ···················· | ···················· | ···················· | ···················· | |
| C | 3080 hCoV-19/Spain/VC-FISABIO-521/2020\|EPI_ISL_447472\|2 | ···················· | ···················· | ···················· | ···················· | ···················· | ···················· | |
| C | 3081 hCoV-19/Spain/VC-FISABIO-522/2020\|EPI_ISL_447473\|2 | ···················· | ···················· | ···················· | ···················· | ···················· | ···················· | |
| C | 3082 hCoV-19/Spain/VC-FISABIO-523/2020\|EPI_ISL_447474\|2 | ···················· | ···················· | ···················· | ···················· | ···················· | ···················· | |
| C | 3083 hCoV-19/Spain/VC-FISABIO-526/2020\|EPI_ISL_447477\|2 | ···················· | ···················· | ···················· | ···················· | ···················· | ···················· | |
| C | 3084 hCoV-19/Spain/VC-FISABIO-529/2020\|EPI_ISL_447478\|2 | ···················· | ···················· | ···················· | ···················· | ···················· | ···················· | |
| C | 3085 hCoV-19/Spain/VC-FISABIO-536/2020\|EPI_ISL_447481\|2 | ···················· | ···················· | ···················· | ···················· | ··N················· | ···················· | |
| C | 3089 hCoV-19/England/OXON-AECF1/2020\|EPI_ISL_448780\|202 | ···················· | ···················· | ···················· | ···················· | ···················· | ···················· | |
| C | 3090 hCoV-19/England/OXON-AED94/2020\|EPI_ISL_448783\|202 | ···················· | ···················· | ···················· | ···················· | ···················· | ···················· | |
| C | 3091 hCoV-19/England/OXON-AEDB2/2020\|EPI_ISL_448784\|202 | ···················· | ···················· | ···················· | ···················· | ···················· | ···················· | |
| C | 3092 hCoV-19/England/OXON-AEDEF/2020\|EPI_ISL_448785\|202 | ···················· | ···················· | ···················· | ···················· | ···················· | ···················· | |
| C | 3093 hCoV-19/England/OXON-AEDFE/2020\|EPI_ISL_448786\|202 | ···················· | ···················· | ···················· | ···················· | ···················· | ···················· | |
| C | 3094 hCoV-19/England/OXON-AEE19/2020\|EPI_ISL_448787\|202 | ···················· | ···················· | ···················· | ···················· | ···················· | ···················· | |
| C | 3095 hCoV-19/England/OXON-AEE55/2020\|EPI_ISL_448788\|202 | ···················· | ···················· | ···················· | ···················· | ···················· | ···················· | |
| C | 3096 hCoV-19/England/OXON-AEE64/2020\|EPI_ISL_448789\|202 | ···················· | ·············A······ | ···················· | ···················· | ···················· | ···················· | |
| C | 3097 hCoV-19/England/OXON-AEF9E/2020\|EPI_ISL_448800\|202 | ···················· | ···················· | ···················· | ···················· | ···················· | ···················· | |
| C | 3098 hCoV-19/England/OXON-AEFBC/2020\|EPI_ISL_448801\|202 | ···················· | ···················· | ···················· | ···················· | ···················· | ···················· | |
| C | 3099 hCoV-19/England/OXON-AEFDA/2020\|EPI_ISL_448802\|202 | ···················· | ···················· | ···················· | ···················· | ···················· | ···················· | |
| C | 3100 hCoV-19/England/OXON-AEFE9/2020\|EPI_ISL_448803\|202 | ···················· | ···················· | ···················· | ···················· | ···················· | ···················· | |
| C | 3101 hCoV-19/England/OXON-AEFF8/2020\|EPI_ISL_448804\|202 | ···················· | ···················· | ···················· | ···················· | ···················· | ···················· | |
| C | 3102 hCoV-19/England/OXON-AF06D/2020\|EPI_ISL_448806\|202 | ···················· | ···················· | ···················· | ···················· | ···················· | ···················· | |
| C | 3103 hCoV-19/England/OXON-AF08B/2020\|EPI_ISL_448807\|202 | ···················· | ···················· | ···················· | ···················· | ···················· | ···················· | |
| C | 3104 hCoV-19/England/OXON-AF09A/2020\|EPI_ISL_448808\|202 | ···················· | ···················· | ···················· | ···················· | ···················· | ···················· | |
| C | 3105 hCoV-19/England/OXON-AF0A9/2020\|EPI_ISL_448809\|202 | ···················· | ···················· | ···················· | ···················· | ···················· | ···················· | |
| C | 3106 hCoV-19/England/OXON-AF0C7/2020\|EPI_ISL_448810\|202 | ···················· | ···················· | ···················· | ···················· | ···················· | ···················· | |
| C | 3107 hCoV-19/England/OXON-AF0F4/2020\|EPI_ISL_448811\|202 | ···················· | ···················· | ···················· | ···················· | ···················· | ···················· | |
| C | 3108 hCoV-19/England/OXON-AF100/2020\|EPI_ISL_448812\|202 | ···················· | ···················· | ···················· | ···················· | ···················· | ···················· | |
| C | 3109 hCoV-19/Denmark/ALAB-HH-116/2020\|EPI_ISL_452000\|20 | ···················· | ···················· | ···················· | ···················· | ···················· | ···················· | |
| C | 3110 hCoV-19/Denmark/ALAB-HH-119/2020\|EPI_ISL_452001\|20 | ···················· | ···················· | ···················· | ···················· | ···················· | ···················· | |
| C | 3111 hCoV-19/Denmark/ALAB-HH-121/2020\|EPI_ISL_452002\|20 | ···················· | ···················· | ···················· | ···················· | ···················· | ···················· | |
| C | 3112 hCoV-19/Denmark/ALAB-HH-167/2020\|EPI_ISL_452037\|20 | ···················· | ···················· | ···················· | ···················· | ···················· | ···················· | |
| C | 3113 hCoV-19/Denmark/ALAB-HH-168/2020\|EPI_ISL_452038\|20 | ···················· | ···················· | ···················· | ···················· | ···················· | ···················· | |
| C | 3114 hCoV-19/Denmark/ALAB-HH-169/2020\|EPI_ISL_452039\|20 | ···················· | ···················· | ···················· | ···················· | ···················· | ···················· | |
| C | 3115 hCoV-19/Denmark/ALAB-HH-173/2020\|EPI_ISL_452042\|20 | ···················· | ···················· | ···················· | ···················· | ···················· | ···················· | |
| C | 3116 hCoV-19/Denmark/ALAB-HH-174/2020\|EPI_ISL_452043\|20 | ···················· | ···················· | ···················· | ···················· | ···················· | ···················· | |
| C | 3117 hCoV-19/Denmark/ALAB-HH-176/2020\|EPI_ISL_452044\|20 | ···················· | ···················· | ···················· | ···················· | ···················· | ···················· | |
| C | 3118 hCoV-19/Denmark/ALAB-HH-177/2020\|EPI_ISL_452045\|20 | ···················· | ···················· | ···················· | ···················· | ···················· | ···················· | |

```
C 3119 hCoV-19/Denmark/ALAB-HH-181/2020|EPI_ISL_452048|20 ···················· ···················· ···················· ···················· ···················· ····················
C 3120 hCoV-19/Denmark/ALAB-HH-183/2020|EPI_ISL_452049|20 ···················· ···················· ···················· ···················· ···················· ····················
C 3121 hCoV-19/Denmark/ALAB-HH-186/2020|EPI_ISL_452050|20 ···················· ···················· ···················· ···················· ···················· ····················
C 3122 hCoV-19/Denmark/ALAB-HH-189/2020|EPI_ISL_452052|20 ···················· ···················· ···················· ···················· ···················· ····················
C 3123 hCoV-19/Denmark/ALAB-HH-190/2020|EPI_ISL_452053|20 ···················· ···················· ···················· ···················· ···················· ····················
C 3124 hCoV-19/Denmark/ALAB-HH-191/2020|EPI_ISL_452054|20 ···················· ···················· ···················· ···················· ···················· ····················
C 3125 hCoV-19/Denmark/ALAB-HH-192/2020|EPI_ISL_452055|20 ···················· ···················· ···················· ···················· ···················· ····················
C 3126 hCoV-19/Denmark/ALAB-HH-193/2020|EPI_ISL_452056|20 ···················· ···················· ···················· ···················· ···················· ····················
C 3127 hCoV-19/Denmark/ALAB-HH-194/2020|EPI_ISL_452057|20 ···················· ···················· ···················· ···················· ···················· ····················
C 3128 hCoV-19/Denmark/ALAB-HH-195/2020|EPI_ISL_452058|20 ···················· ···················· ···················· ···················· ···················· ····················
C 3129 hCoV-19/Denmark/ALAB-HH-198/2020|EPI_ISL_452059|20 ···················· ···················· ···················· ···················· ···················· ····················
C 3130 hCoV-19/Denmark/ALAB-HH-199/2020|EPI_ISL_452060|20 ···················· ···················· ···················· ···················· ···················· ····················
C 3131 hCoV-19/Denmark/ALAB-HH-204/2020|EPI_ISL_452061|20 ···················· ···················· ···················· ···················· ···················· ····················
C 3132 hCoV-19/Denmark/ALAB-HH-206/2020|EPI_ISL_452062|20 ···················· ···················· ···················· ···················· ···················· ····················
C 3133 hCoV-19/Denmark/ALAB-HH-209/2020|EPI_ISL_452064|20 ···················· ···················· ···················· ···················· ···················· ····················
C 3134 hCoV-19/Denmark/ALAB-HH-210/2020|EPI_ISL_452065|20 ···················· ···················· ···················· ···················· ···················· ····················
C 3135 hCoV-19/Denmark/ALAB-HH-211/2020|EPI_ISL_452066|20 ···················· ···················· ···················· ···················· ···················· ····················
C 3136 hCoV-19/Denmark/ALAB-HH-212/2020|EPI_ISL_452067|20 ···················· ···················· ···················· ···················· ···················· ····················
C 3137 hCoV-19/Denmark/ALAB-HH-213/2020|EPI_ISL_452068|20 ···················· ···················· ···················· ···················· ···················· ····················
C 3138 hCoV-19/Denmark/ALAB-HH-218/2020|EPI_ISL_452069|20 ···················· ···················· ···················· ···················· ···················· ····················
C 3139 hCoV-19/Denmark/ALAB-HH-219/2020|EPI_ISL_452070|20 ···················· ···················· ···················· ···················· ···················· ····················
C 3140 hCoV-19/Denmark/ALAB-HH-224/2020|EPI_ISL_452074|20 ···················· ···················· ···················· ···················· ···················· ····················
C 3141 hCoV-19/Denmark/ALAB-HH-227/2020|EPI_ISL_452075|20 ···················· ···················· ···················· ···················· ···················· ····················
C 3142 hCoV-19/Denmark/ALAB-HH-228/2020|EPI_ISL_452076|20 ···················· ···················· ···················· ···················· ···················· ····················
C 3143 hCoV-19/Denmark/ALAB-HH-233/2020|EPI_ISL_452077|20 ···················· ···················· ···················· ···················· ···················· ····················
C 3144 hCoV-19/Denmark/ALAB-HH-235/2020|EPI_ISL_452078|20 ···················· ···················· ···················· ···················· ···················· ····················
C 3145 hCoV-19/Denmark/ALAB-HH-238/2020|EPI_ISL_452079|20 ···················· ···················· ···················· ···················· ···················· ····················
C 3146 hCoV-19/Kazakhstan/26565/2020|EPI_ISL_454600|2020- ···················· ···················· ···················· ···················· ···················· ····················
C 3147 hCoV-19/Kazakhstan/26568/2020|EPI_ISL_454601|2020- ···················· ···················· ···················· ···················· ···················· ····················
C 3148 hCoV-19/Croatia/14412_Koprivnica/2020|EPI_ISL_4546 ···················· ···················· ···················· ···················· ···················· ····················
C 3149 hCoV-19/Kazakhstan/26579/2020|EPI_ISL_454603|2020- ···················· ···················· ···················· ···················· ···················· ····················
C 3150 hCoV-19/Kazakhstan/26580/2020|EPI_ISL_454604|2020- ···················· ···················· ···················· ···················· ···················· ····················
C 3151 hCoV-19/Croatia/8U-S17new/2020|EPI_ISL_454605|2020 ···················· ···················· ···················· ···················· ···················· ····················
C 3152 hCoV-19/Croatia/I7-S21new/2020|EPI_ISL_454606|2020 ···················· ···················· ···················· ···················· ···················· ····················
C 3153 hCoV-19/USA/CA-CZB-1270/2020|EPI_ISL_454607|2020-0 ···················· ···················· ···················· ···················· ···················· ····················
C 3154 hCoV-19/USA/CA-CZB-1271/2020|EPI_ISL_454608|2020-0 ···················· ···················· ···················· ···················· ···················· ····················
C 3155 hCoV-19/USA/CA-CZB-1272/2020|EPI_ISL_454609|2020-0 ···················· ···················· ···················· ···················· ···················· ····················
C 3156 hCoV-19/USA/CA-CZB-1273/2020|EPI_ISL_454610|2020-0 ···················· ···················· ···················· ···················· ···················· ····················
C 3157 hCoV-19/USA/CA-CZB-1274/2020|EPI_ISL_454611|2020-0 ···················· ···················· ···················· ···················· ···················· ····················
C 3158 hCoV-19/USA/CA-CZB-1275/2020|EPI_ISL_454612|2020-0 ···················· ···················· ···················· ···················· ···················· ····················
C 3159 hCoV-19/USA/CA-CZB-1276/2020|EPI_ISL_454613|2020-0 ···················· ···················· ···················· ···················· ···················· ····················
C 3160 hCoV-19/USA/CA-CZB-1150/2020|EPI_ISL_454615|2020-0 ···················· ···················· ···················· ···················· ···················· ····················
C 3161 hCoV-19/USA/CA-CZB-1156/2020|EPI_ISL_454617|2020-0 ···················· ···················· ···················· ···················· ···················· ····················
C 3162 hCoV-19/USA/CA-CZB-1157/2020|EPI_ISL_454618|2020-0 ···················· ···················· ···················· ···················· ···················· ····················
C 3163 hCoV-19/USA/CA-CZB-1160/2020|EPI_ISL_454619|2020-0 ···················· ···················· ···················· ···················· ···················· ····················
C 3164 hCoV-19/USA/CA-CZB-1163/2020|EPI_ISL_454620|2020-0 ···················· ···················· ···················· ···················· ···················· ····················
C 3165 hCoV-19/USA/CA-CZB-1164/2020|EPI_ISL_454621|2020-0 ···················· ···················· ···················· ···················· ···················· ····················
C 3166 hCoV-19/USA/CA-CZB-1168/2020|EPI_ISL_454622|2020-0 ···················· ···················· ···················· ···················· ···················· ····················
C 3167 hCoV-19/USA/CA-CZB-1171/2020|EPI_ISL_454624|2020-0 ···················· ···················· ···················· ···················· ···················· ····················
C 3168 hCoV-19/USA/CA-CZB-1174/2020|EPI_ISL_454625|2020-0 ···················· ···················· ···················· ···················· ···················· ····················
C 3169 hCoV-19/USA/CA-CZB-1175/2020|EPI_ISL_454626|2020-0 ···················· ···················· ···················· ···················· ···················· ····················
C 3170 hCoV-19/USA/CA-CZB-1176/2020|EPI_ISL_454627|2020-0 ···················· ···················· ···················· ···················· ···················· ····················
C 3171 hCoV-19/USA/CA-CZB-1178/2020|EPI_ISL_454628|2020-0 ···················· ···················· ···················· ···················· ···················· ····················
C 3172 hCoV-19/USA/CA-CZB-1179/2020|EPI_ISL_454629|2020-0 ···················· ···················· ···················· ···················· ···················· ····················
C 3173 hCoV-19/USA/CA-CZB-1183/2020|EPI_ISL_454630|2020-0 ···················· ···················· ···················· ···················· ···················· ····················
C 3174 hCoV-19/USA/CA-CZB-1186/2020|EPI_ISL_454631|2020-0 ···················· ···················· ···················· ···················· ···················· ····················
C 3175 hCoV-19/USA/CA-CZB-1189/2020|EPI_ISL_454632|2020-0 ···················· ···················· ···················· ···················· ···················· ····················
C 3176 hCoV-19/USA/CA-CZB-1193/2020|EPI_ISL_454633|2020-0 ···················· ···················· ···················· ···················· ···················· ····················
C 3177 hCoV-19/USA/CA-CZB-1195/2020|EPI_ISL_454634|2020-0 ···················· ···················· ···················· ···················· ···················· ····················
C 3178 hCoV-19/USA/CA-CZB-1277/2020|EPI_ISL_454635|2020-0 ···················· ···················· ···················· ···················· ···················· ····················
C 3179 hCoV-19/USA/CA-CZB-1278/2020|EPI_ISL_454636|2020-0 ···················· ···················· ···················· ···················· ···················· ····················
C 3180 hCoV-19/USA/CA-CZB-1279/2020|EPI_ISL_454637|2020-0 ···················· ···················· ···················· ···················· ···················· ····················
C 3181 hCoV-19/USA/CA-CZB-1280/2020|EPI_ISL_454638|2020-0 ···················· ···················· ···················· ···················· ···················· ····················
C 3182 hCoV-19/USA/CA-CZB-1281/2020|EPI_ISL_454639|2020-0 ···················· ···················· ···················· ···················· ···················· ····················
C 3183 hCoV-19/USA/CA-CZB-1282/2020|EPI_ISL_454640|2020-0 ···················· ···················· ···················· ···················· ···················· ····················
C 3184 hCoV-19/USA/CA-CZB-1283/2020|EPI_ISL_454641|2020-0 ···················· ···················· ···················· ···················· ···················· ····················
C 3185 hCoV-19/USA/CT-CDC-6001/2020|EPI_ISL_454642|2020-0 ···················· ···················· ···················· ···················· ···················· ····················
C 3186 hCoV-19/USA/VI-CDC-4022/2020|EPI_ISL_454643|2020-0 ···················· ···················· ···················· ···················· ···················· ····················
C 3187 hCoV-19/USA/VI-CDC-4411/2020|EPI_ISL_454644|2020-0 ···················· ···················· ···················· ···················· ···················· ····················
C 3188 hCoV-19/USA/CT-CDC-8001/2020|EPI_ISL_454645|2020-0 ···················· ···················· ···················· ···················· ···················· ····················
C 3189 hCoV-19/USA/TX-CDC-0406/2020|EPI_ISL_454646|2020-0 ···················· ···················· ···················· ···················· ···················· ····················
C 3190 hCoV-19/USA/VI-CDC-4399/2020|EPI_ISL_454647|2020-0 ···················· ···················· ···················· ···················· ···················· ····················
C 3191 hCoV-19/USA/VI-CDC-4400/2020|EPI_ISL_454648|2020-0 ···················· ···················· ···················· ···················· ···················· ····················
C 3192 hCoV-19/USA/VI-CDC-4371/2020|EPI_ISL_454649|2020-0 ···················· ···················· ···················· ···················· ···················· ····················
C 3193 hCoV-19/USA/VI-CDC-4367/2020|EPI_ISL_454650|2020-0 ···················· ···················· ···················· ···················· ···················· ····················
C 3194 hCoV-19/USA/VI-CDC-4408/2020|EPI_ISL_454651|2020-0 ···················· ···················· ···················· ···················· ···················· ····················
C 3195 hCoV-19/USA/VI-CDC-4413/2020|EPI_ISL_454652|2020-0 ···················· ···················· ···················· ···················· ···················· ····················
C 3196 hCoV-19/USA/CA-CZB-1241/2020|EPI_ISL_454661|2020-0 ···················· ···················· ···················· ···················· ···················· ····················
```

| | | | | | | | | |
|---|---|---|---|---|---|---|---|---|
| C | 3197 | hCoV-19/USA/CA-CZB-1242/2020\|EPI_ISL_454662\|2020-0 | ···················· | ···················· | ···················· | ···················· | ···················· | ···················· |
| C | 3198 | hCoV-19/USA/CA-CZB-1243/2020\|EPI_ISL_454663\|2020-0 | ···················· | ···················· | ···················· | ···················· | ···················· | ···················· |
| C | 3199 | hCoV-19/USA/CA-CZB-1244/2020\|EPI_ISL_454664\|2020-0 | ···················· | ···················· | ···················· | ···················· | ···················· | ···················· |
| C | 3200 | hCoV-19/Sweden/20-50287/2020\|EPI_ISL_455900\|2020-0 | ···················· | ···················· | ···················· | ···················· | ···················· | ···················· |
| C | 3201 | hCoV-19/Sweden/20-50289/2020\|EPI_ISL_455901\|2020-0 | ···················· | ···················· | ···················· | ···················· | ···················· | ···················· |
| C | 3202 | hCoV-19/Sweden/20-08127/2020\|EPI_ISL_455902\|2020-0 | ···················· | ···················· | ···················· | ···················· | ···················· | ···················· |
| C | 3203 | hCoV-19/Sweden/20-50279/2020\|EPI_ISL_455903\|2020-0 | ···················· | ···················· | ···················· | ···················· | ···················· | ···················· |
| C | 3204 | hCoV-19/Sweden/20-50270/2020\|EPI_ISL_455904\|2020-0 | ···················· | ···················· | ···················· | ···················· | ···················· | ···················· |
| C | 3205 | hCoV-19/Sweden/20-50272/2020\|EPI_ISL_455905\|2020-0 | ···················· | ···················· | ···················· | ···················· | ···················· | ···················· |
| C | 3206 | hCoV-19/Sweden/20-50274/2020\|EPI_ISL_455906\|2020-0 | ···················· | ···················· | ···················· | ···················· | ···················· | ···················· |
| C | 3207 | hCoV-19/Sweden/20-50266/2020\|EPI_ISL_455907\|2020-0 | ···················· | ···················· | ···················· | ···················· | ···················· | ···················· |
| C | 3208 | hCoV-19/Thailand/Bangkok-CONI-0106/2020\|EPI_ISL_45 | ···················· | ···················· | ···················· | ···················· | ···················· | ···················· |
| C | 3209 | hCoV-19/Thailand/Bangkok-CONI-0107/2020\|EPI_ISL_45 | ···················· | ···················· | ···················· | ···················· | ···················· | ···················· |
| C | 3210 | hCoV-19/Thailand/Bangkok-CONI-0108/2020\|EPI_ISL_45 | ···················· | ···················· | ···················· | ···················· | ···················· | ···················· |
| C | 3212 | hCoV-19/Thailand/Bangkok-CONI-0110/2020\|EPI_ISL_45 | ···················· | ······C············· | ···················· | ···················· | ···················· | ···················· |
| C | 3214 | hCoV-19/Thailand/Bangkok-CONI-0115/2020\|EPI_ISL_45 | ···················· | ···················· | ···················· | ···················· | ···················· | ···················· |
| C | 3215 | hCoV-19/Thailand/Bangkok-CONI-0128/2020\|EPI_ISL_45 | ···················· | ···················· | ···················· | ···················· | ···················· | ···················· |
| C | 3216 | hCoV-19/Thailand/Bangkok-CONI-0131/2020\|EPI_ISL_45 | ···················· | ···················· | ···················· | ···················· | ···················· | ···················· |
| C | 3217 | hCoV-19/Thailand/Bangkok-CONI-0132/2020\|EPI_ISL_45 | ···················· | ···················· | ···················· | ···················· | ···················· | ···················· |
| C | 3218 | hCoV-19/Thailand/Bangkok-CONI-0133/2020\|EPI_ISL_45 | ···················· | ···················· | ···················· | ···················· | ···················· | ···················· |
| C | 3219 | hCoV-19/Thailand/Bangkok-CONI-0134/2020\|EPI_ISL_45 | ···················· | ···················· | ···················· | ···················· | ···················· | ···················· |
| C | 3220 | hCoV-19/Thailand/Bangkok-CONI-0137/2020\|EPI_ISL_45 | ···················· | ···················· | ···················· | ···················· | ···················· | ···················· |
| C | 3221 | hCoV-19/Thailand/Bangkok-CONI-0138/2020\|EPI_ISL_45 | ···················· | ···················· | ···················· | ···················· | ···················· | ···················· |
| C | 3222 | hCoV-19/Thailand/Bangkok-CONI-0140/2020\|EPI_ISL_45 | ···················· | ···················· | ···················· | ···················· | ···················· | ···················· |
| C | 3223 | hCoV-19/Thailand/Bangkok-CONI-0141/2020\|EPI_ISL_45 | ···················· | ···················· | ···················· | ···················· | ···················· | ···················· |
| C | 3224 | hCoV-19/Thailand/Bangkok-CONI-0142/2020\|EPI_ISL_45 | ···················· | ···················· | ···················· | ···················· | ···················· | ···················· |
| C | 3225 | hCoV-19/Thailand/Bangkok-CONI-0144/2020\|EPI_ISL_45 | ···················· | ···················· | ···················· | ···················· | ···················· | ···················· |
| C | 3226 | hCoV-19/Thailand/Bangkok-CONI-0145/2020\|EPI_ISL_45 | ···················· | ···················· | ···················· | ···················· | ···················· | ···················· |
| C | 3227 | hCoV-19/Thailand/Bangkok-CONI-0146/2020\|EPI_ISL_45 | ···················· | ···················· | ···················· | ···················· | ···················· | ···················· |
| C | 3228 | hCoV-19/Thailand/Bangkok-CONI-0147/2020\|EPI_ISL_45 | ···················· | ···················· | ···················· | ···················· | ···················· | ···················· |
| C | 3229 | hCoV-19/Thailand/Bangkok-CONI-0148/2020\|EPI_ISL_45 | ···················· | ···················· | ···················· | ···················· | ···················· | ···················· |
| C | 3231 | hCoV-19/Thailand/Bangkok-CONI-0154/2020\|EPI_ISL_45 | ···················· | ···················· | ···················· | ···················· | ···················· | ···················· |
| C | 3232 | hCoV-19/Belgium/ULG-10170/2020\|EPI_ISL_455958\|2020 | ···················· | ···················· | ···················· | ···················· | ···················· | ···················· |
| C | 3233 | hCoV-19/Belgium/ULG-10173/2020\|EPI_ISL_455961\|2020 | ···················· | ···················· | ···················· | ···················· | ···················· | ···················· |
| C | 3234 | hCoV-19/Belgium/ULG-10174/2020\|EPI_ISL_455962\|2020 | ···················· | ···················· | ···················· | ···················· | ···················· | ···················· |
| C | 3235 | hCoV-19/Belgium/ULG-10175/2020\|EPI_ISL_455963\|2020 | ···················· | ···················· | ···················· | ···················· | ···················· | ···················· |
| C | 3236 | hCoV-19/Belgium/ULG-10177/2020\|EPI_ISL_455965\|2020 | ···················· | ···················· | ···················· | ···················· | ···················· | ···················· |
| C | 3237 | hCoV-19/Belgium/ULG-10178/2020\|EPI_ISL_455966\|2020 | ···················· | ···················· | ···················· | ···················· | ···················· | ···················· |
| C | 3238 | hCoV-19/Belgium/ULG-10179/2020\|EPI_ISL_455967\|2020 | ···················· | ···················· | ···················· | ···················· | ···················· | ···················· |
| C | 3239 | hCoV-19/Belgium/ULG-10180/2020\|EPI_ISL_455968\|2020 | ···················· | ···················· | ···················· | ···················· | ···················· | ···················· |
| C | 3240 | hCoV-19/Belgium/ULG-10181/2020\|EPI_ISL_455969\|2020 | ···················· | ···················· | ···················· | ···················· | ···················· | ···················· |
| C | 3241 | hCoV-19/Belgium/ULG-10182/2020\|EPI_ISL_455970\|2020 | ···················· | ···················· | ···················· | ···················· | ···················· | ···················· |
| C | 3242 | hCoV-19/Belgium/ULG-10184/2020\|EPI_ISL_455972\|2020 | ···················· | ···················· | ···················· | ···················· | ···················· | ···················· |
| C | 3243 | hCoV-19/Belgium/ULG-10185/2020\|EPI_ISL_455973\|2020 | ···················· | ···················· | ···················· | ···················· | ···················· | ···················· |
| C | 3244 | hCoV-19/Belgium/ULG-10186/2020\|EPI_ISL_455974\|2020 | ···················· | ···················· | ···················· | ···················· | ···················· | ···················· |
| C | 3245 | hCoV-19/Belgium/ULG-10187/2020\|EPI_ISL_455975\|2020 | ···················· | ···················· | ···················· | ···················· | ···················· | ···················· |
| C | 3246 | hCoV-19/Belgium/ULG-10188/2020\|EPI_ISL_455976\|2020 | ···················· | ···················· | ···················· | ···················· | ···················· | ···················· |
| C | 3247 | hCoV-19/Belgium/ULG-10189/2020\|EPI_ISL_455977\|2020 | ···················· | ···················· | ···················· | ···················· | ···················· | ···················· |
| C | 3248 | hCoV-19/Belgium/ULG-10190/2020\|EPI_ISL_455978\|2020 | ···················· | ···················· | ···················· | ···················· | ···················· | ···················· |
| C | 3249 | hCoV-19/Belgium/ULG-10191/2020\|EPI_ISL_455979\|2020 | ···················· | ···················· | ···················· | ···················· | ···················· | ···················· |
| C | 3250 | hCoV-19/USA/LA-EVTL137/2020\|EPI_ISL_455980\|2020-03 | ···················· | ···················· | ···················· | ···················· | ···················· | ···················· |
| C | 3251 | hCoV-19/USA/LA-EVTL138/2020\|EPI_ISL_455981\|2020-03 | ···················· | ···················· | ···················· | ···················· | ···················· | ···················· |
| C | 3252 | hCoV-19/USA/LA-EVTL139/2020\|EPI_ISL_455982\|2020-03 | ···················· | ···················· | ···················· | ···················· | ···················· | ···················· |
| C | 3253 | hCoV-19/USA/LA-EVTL140/2020\|EPI_ISL_455983\|2020-03 | ···················· | ···················· | ···················· | ···················· | ···················· | ···················· |
| C | 3254 | hCoV-19/USA/LA-EVTL141/2020\|EPI_ISL_455984\|2020-03 | ···················· | ···················· | ···················· | ···················· | ···················· | ···················· |
| C | 3255 | hCoV-19/USA/TX-EVTL149/2020\|EPI_ISL_455992\|2020-03 | ···················· | ···················· | ···················· | ···················· | ···················· | ···················· |
| C | 3256 | hCoV-19/USA/LA-EVTL150/2020\|EPI_ISL_455993\|2020-03 | ···················· | ···················· | ···················· | ···················· | ···················· | ···················· |
| C | 3257 | hCoV-19/USA/LA-EVTL151/2020\|EPI_ISL_455994\|2020-04 | ···················· | ···················· | ···················· | ···················· | ···················· | ···················· |
| C | 3258 | hCoV-19/USA/LA-EVTL152/2020\|EPI_ISL_455995\|2020-04 | ···················· | ···················· | ···················· | ···················· | ···················· | ···················· |
| C | 3259 | hCoV-19/India/GJ-GBRC148a/2020\|EPI_ISL_458109\|2020 | ···················· | ···················· | ···················· | ···················· | ···················· | ···················· |
| C | 3260 | hCoV-19/Oman/205028472/2020\|EPI_ISL_458119\|2020-04 | ···················· | ···················· | ···················· | ···················· | ···················· | ···················· |
| C | 3261 | hCoV-19/Oman/205028997/2020\|EPI_ISL_458120\|2020-04 | ···················· | ···················· | ···················· | ···················· | ···················· | ···················· |
| C | 3262 | hCoV-19/Oman/205030448/2020\|EPI_ISL_458121\|2020-04 | ···················· | ···················· | ···················· | ···················· | ···················· | ···················· |
| C | 3263 | hCoV-19/Oman/205027285/2020\|EPI_ISL_458122\|2020-04 | ···················· | ···················· | ···················· | ···················· | ···················· | ···················· |
| C | 3264 | hCoV-19/Oman/205027055/2020\|EPI_ISL_458123\|2020-04 | ···················· | ···················· | ···················· | ···················· | ···················· | ···················· |
| C | 3265 | hCoV-19/Oman/205033391/2020\|EPI_ISL_458124\|2020-05 | ···················· | ···················· | ···················· | ···················· | ···················· | ···················· |
| C | 3266 | hCoV-19/Spain/CT-HUVH-VH8002/2020\|EPI_ISL_458130\|2 | ···················· | ···················· | ···················· | ···················· | ···················· | ···················· |
| C | 3267 | hCoV-19/Spain/CT-HUVH-VH9461/2020\|EPI_ISL_458131\|2 | ···················· | ···················· | ···················· | ···················· | ···················· | ···················· |
| C | 3268 | hCoV-19/Spain/CT-HUVH-VH9758/2020\|EPI_ISL_458132\|2 | ···················· | ···················· | ···················· | ···················· | ···················· | ···················· |
| C | 3269 | hCoV-19/Bangladesh/NIB-BCSIR-02/2020\|EPI_ISL_45813 | ···················· | ···················· | ···················· | ···················· | ···················· | ···················· |
| C | 3270 | hCoV-19/Brazil/AP-IEC-162741/2020\|EPI_ISL_458138\|2 | ···················· | ···················· | ···················· | ···················· | ···················· | ···················· |
| C | 3271 | hCoV-19/Brazil/AC-IEC-162535/2020\|EPI_ISL_458139\|2 | ···················· | ···················· | ···················· | ···················· | ···················· | ···················· |
| C | 3272 | hCoV-19/Belgium/Rega-0327210/2020\|EPI_ISL_458180\|2 | ···················· | ···················· | ···················· | ···················· | ···················· | ···················· |
| C | 3273 | hCoV-19/Belgium/Rega-0327211/2020\|EPI_ISL_458181\|2 | ···················· | ···················· | ···················· | ···················· | ···················· | ···················· |
| C | 3274 | hCoV-19/Belgium/Rega-0327212/2020\|EPI_ISL_458182\|2 | ···················· | ···················· | ···················· | ···················· | ···················· | ···················· |
| C | 3275 | hCoV-19/Belgium/Rega-0327213/2020\|EPI_ISL_458183\|2 | ···················· | ···················· | ···················· | ···················· | ···················· | ···················· |
| C | 3276 | hCoV-19/Belgium/Rega-0327214/2020\|EPI_ISL_458184\|2 | ···················· | ···················· | ···················· | ···················· | ···················· | ···················· |
| C | 3277 | hCoV-19/Belgium/Rega-0327215/2020\|EPI_ISL_458185\|2 | ···················· | ···················· | ···················· | ···················· | ···················· | ···················· |

| | | | | | | | |
|---|---|---|---|---|---|---|---|
| C | 3278 hCoV-19/Belgium/Rega-0327216/2020\|EPI_ISL_458186\|2 | ···················· | ···················· | ···················· | ···················· | ···················· | ···················· |
| C | 3279 hCoV-19/Belgium/Rega-0329217/2020\|EPI_ISL_458187\|2 | ···················· | ···················· | ···················· | ···················· | ···················· | ···················· |
| C | 3280 hCoV-19/Belgium/Rega-0329218/2020\|EPI_ISL_458188\|2 | ···················· | ···················· | ···················· | ···················· | ···················· | ···················· |
| C | 3281 hCoV-19/Belgium/Rega-0329219/2020\|EPI_ISL_458189\|2 | ···················· | ···················· | ···················· | ···················· | ···················· | ···················· |
| C | 3282 hCoV-19/Belgium/Rega-0329220/2020\|EPI_ISL_458190\|2 | ···················· | ···················· | ···················· | ···················· | ···················· | ···················· |
| C | 3283 hCoV-19/Belgium/Rega-0329221/2020\|EPI_ISL_458191\|2 | ···················· | ···················· | ···················· | ···················· | ···················· | ···················· |
| C | 3284 hCoV-19/Belgium/Rega-0329222/2020\|EPI_ISL_458192\|2 | ···················· | ···················· | ···················· | ···················· | ···················· | ···················· |
| C | 3285 hCoV-19/Belgium/Rega-0329223/2020\|EPI_ISL_458193\|2 | ···················· | ···················· | ···················· | ···················· | ···················· | ···················· |
| C | 3286 hCoV-19/Belgium/Rega-0329224/2020\|EPI_ISL_458194\|2 | ···················· | ···················· | ···················· | ···················· | ···················· | ···················· |
| C | 3287 hCoV-19/Belgium/Rega-0329225/2020\|EPI_ISL_458195\|2 | ···················· | ···················· | ···················· | ···················· | ···················· | ···················· |
| C | 3288 hCoV-19/Belgium/Rega-0329226/2020\|EPI_ISL_458196\|2 | ···················· | ···················· | ···················· | ···················· | ···················· | ···················· |
| C | 3289 hCoV-19/Belgium/Rega-0329227/2020\|EPI_ISL_458197\|2 | ···················· | ···················· | ···················· | ···················· | ···················· | ···················· |
| C | 3290 hCoV-19/Belgium/Rega-0329228/2020\|EPI_ISL_458198\|2 | ···················· | ···················· | ···················· | ···················· | ···················· | ···················· |
| C | 3291 hCoV-19/Belgium/Rega-0329229/2020\|EPI_ISL_458199\|2 | ···················· | ···················· | ···················· | ···················· | ···················· | ···················· |
| C | 3292 hCoV-19/England/CAMB-1ADE0A/2020\|EPI_ISL_459453\|20 | ···················· | ···················· | ···················· | ···················· | ···················· | ···················· |
| C | 3293 hCoV-19/England/CAMB-1AE23A/2020\|EPI_ISL_459455\|20 | ···················· | ···················· | ···················· | ···················· | ···················· | ···················· |
| C | 3294 hCoV-19/England/CAMB-1AE4BC/2020\|EPI_ISL_459456\|20 | ···················· | ···················· | ···················· | ···················· | ···················· | ···················· |
| C | 3295 hCoV-19/England/CAMB-1ADD94/2020\|EPI_ISL_459459\|20 | ···················· | ···················· | ···················· | ···················· | ···················· | ···················· |
| C | 3296 hCoV-19/England/CAMB-1AE197/2020\|EPI_ISL_459461\|20 | ···················· | ···················· | ···················· | ···················· | ···················· | ···················· |
| C | 3297 hCoV-19/England/CAMB-1ADFE9/2020\|EPI_ISL_459463\|20 | ···················· | ···················· | ···················· | ···················· | ···················· | ···················· |
| C | 3298 hCoV-19/England/CAMB-1AE05E/2020\|EPI_ISL_459466\|20 | ···················· | ···················· | ···················· | ···················· | ···················· | ···················· |
| C | 3299 hCoV-19/England/CAMB-1AE64D/2020\|EPI_ISL_459467\|20 | ···················· | ···················· | ···················· | ···················· | ···················· | ···················· |
| C | 3300 hCoV-19/England/CAMB-1ADDEF/2020\|EPI_ISL_459468\|20 | ···················· | ···················· | ···················· | ···················· | ···················· | ···················· |
| C | 3301 hCoV-19/USA/MA-MGH-00304/2020\|EPI_ISL_460430\|2020- | ···················· | ···················· | ···················· | ···················· | ···················· | ···················· |
| C | 3302 hCoV-19/USA/MA-MGH-00307/2020\|EPI_ISL_460431\|2020- | ···················· | ···················· | ···················· | ···················· | ···················· | ···················· |
| C | 3303 hCoV-19/USA/MA-MGH-00305/2020\|EPI_ISL_460432\|2020- | ···················· | ···················· | ···················· | ···················· | ···················· | ···················· |
| C | 3304 hCoV-19/USA/MA-MGH-00439/2020\|EPI_ISL_460434\|2020- | ···················· | ···················· | ···················· | ···················· | ···················· | ···················· |
| C | 3305 hCoV-19/USA/MA-MGH-00245/2020\|EPI_ISL_460435\|2020- | ···················· | ···················· | ···················· | ···················· | ···················· | ···················· |
| C | 3306 hCoV-19/USA/MA-MGH-00531/2020\|EPI_ISL_460436\|2020- | ···················· | ···················· | ···················· | ···················· | ···················· | ···················· |
| C | 3307 hCoV-19/USA/MA-MGH-00202/2020\|EPI_ISL_460437\|2020- | ···················· | ···················· | ···················· | ···················· | ···················· | ···················· |
| C | 3308 hCoV-19/USA/MA-MGH-00006/2020\|EPI_ISL_460440\|2020- | ···················· | ···················· | ···················· | ···················· | ···················· | ···················· |
| C | 3309 hCoV-19/USA/MA-MGH-00481/2020\|EPI_ISL_460441\|2020- | ···················· | ···················· | ···················· | ···················· | ···················· | ···················· |
| C | 3310 hCoV-19/USA/MA-MGH-00432/2020\|EPI_ISL_460442\|2020- | ···················· | ···················· | ···················· | ···················· | ···················· | ···················· |
| C | 3311 hCoV-19/USA/MA-MGH-00635/2020\|EPI_ISL_460443\|2020- | ···················· | ···················· | ···················· | ···················· | ···················· | ···················· |
| C | 3312 hCoV-19/USA/MA-MGH-00084/2020\|EPI_ISL_460444\|2020- | ···················· | ···················· | ···················· | ···················· | ···················· | ···················· |
| C | 3313 hCoV-19/USA/MA-MGH-00505/2020\|EPI_ISL_460445\|2020- | ···················· | ···················· | ···················· | ···················· | ···················· | ···················· |
| C | 3314 hCoV-19/USA/MA-MGH-00071/2020\|EPI_ISL_460446\|2020- | ···················· | ···················· | ···················· | ···················· | ···················· | ···················· |
| C | 3315 hCoV-19/USA/MA-MGH-00152/2020\|EPI_ISL_460447\|2020- | ···················· | ···················· | ···················· | ···················· | ···················· | ···················· |
| C | 3316 hCoV-19/USA/MA-MGH-00075/2020\|EPI_ISL_460448\|2020- | ···················· | ···················· | ···················· | ···················· | ···················· | ···················· |
| C | 3317 hCoV-19/USA/MA-MGH-00233/2020\|EPI_ISL_460450\|2020- | ···················· | ···················· | ···················· | ···················· | ···················· | ···················· |
| C | 3318 hCoV-19/Scotland/EDB608/2020\|EPI_ISL_461761\|2020-0 | ···················· | ···················· | ···················· | ···················· | ···················· | ···················· |
| C | 3319 hCoV-19/Scotland/EDB817/2020\|EPI_ISL_461762\|2020-0 | ···················· | ···················· | ···················· | ···················· | ···················· | ···················· |
| C | 3320 hCoV-19/England/LOND-D6A41/2020\|EPI_ISL_461763\|202 | ···················· | ···················· | ···················· | ···················· | ···················· | ···················· |
| C | 3321 hCoV-19/England/LOND-D6A50/2020\|EPI_ISL_461764\|202 | ···················· | ···················· | ···················· | ···················· | ···················· | ···················· |
| C | 3322 hCoV-19/England/LOND-D6A6F/2020\|EPI_ISL_461765\|202 | ···················· | ···················· | ···················· | ···················· | ···················· | ···················· |
| C | 3323 hCoV-19/England/LOND-D6A7E/2020\|EPI_ISL_461766\|202 | ···················· | ···················· | ···················· | ···················· | ···················· | ···················· |
| C | 3324 hCoV-19/England/LOND-D6A8D/2020\|EPI_ISL_461767\|202 | ···················· | ···················· | ···················· | ···················· | ···················· | ···················· |
| C | 3325 hCoV-19/England/LOND-D6A9C/2020\|EPI_ISL_461768\|202 | ···················· | ···················· | ···················· | ···················· | ···················· | ···················· |
| C | 3326 hCoV-19/England/LOND-D6ABA/2020\|EPI_ISL_461769\|202 | ···················· | ···················· | ···················· | ···················· | ···················· | ···················· |
| C | 3327 hCoV-19/England/LOND-D6AC9/2020\|EPI_ISL_461770\|202 | ···················· | ···················· | ···················· | ···················· | ···················· | ···················· |
| C | 3328 hCoV-19/England/LOND-D6AE7/2020\|EPI_ISL_461771\|202 | ···················· | ···················· | ···················· | ···················· | ···················· | ···················· |
| C | 3330 hCoV-19/Israel/2046616/2020\|EPI_ISL_447401\|2020-04 | ···················· | ···················· | ···················· | ···················· | ···················· | ·············N······ |
| C | 3331 hCoV-19/Israel/2046614/2020\|EPI_ISL_447402\|2020-04 | ···················· | ···················· | ···················· | ···················· | ···················· | ···················· |
| C | 3332 hCoV-19/Israel/2046548/2020\|EPI_ISL_447403\|2020-04 | ···················· | ···················· | ···················· | ···················· | ···················· | ···················· |
| C | 3333 hCoV-19/Israel/2046434/2020\|EPI_ISL_447404\|2020-04 | ···················· | ···················· | ···················· | ···················· | ···················· | ···················· |
| C | 3334 hCoV-19/Israel/13077723/2020\|EPI_ISL_447427\|2020-0 | ···················· | ···················· | ···················· | ···················· | ···················· | ···················· |
| C | 3335 hCoV-19/Israel/13077711/2020\|EPI_ISL_447428\|2020-0 | ···················· | ···················· | ···················· | ···················· | ···················· | ···················· |
| C | 3336 hCoV-19/Israel/13077564/2020\|EPI_ISL_447429\|2020-0 | ···················· | ···················· | ···················· | ···················· | ···················· | ···················· |
| C | 3338 hCoV-19/Israel/13077558/2020\|EPI_ISL_447432\|2020-0 | ···················· | ···················· | ···················· | ···················· | ···················· | ···················· |
| C | 3339 hCoV-19/Israel/13077510/2020\|EPI_ISL_447434\|2020-0 | ···················· | ···················· | ···················· | ···················· | ···················· | ···················· |
| C | 3340 hCoV-19/Israel/13077498/2020\|EPI_ISL_447435\|2020-0 | ···················· | ···················· | ···················· | ···················· | ···················· | ···················· |
| C | 3341 hCoV-19/Israel/13077497/2020\|EPI_ISL_447436\|2020-0 | ···················· | ···················· | ···················· | ···················· | ···················· | ···················· |
| C | 3342 hCoV-19/Israel/13077413/2020\|EPI_ISL_447438\|2020-0 | ···················· | ···················· | ···················· | ···················· | ···················· | ···················· |
| C | 3343 hCoV-19/Israel/13077377/2020\|EPI_ISL_447440\|2020-0 | ···················· | ···················· | ···················· | ···················· | ···················· | ···················· |
| C | 3344 hCoV-19/Israel/13075914/2020\|EPI_ISL_447441\|2020-0 | ···················· | ···················· | ···················· | ···················· | ···················· | ···················· |
| C | 3345 hCoV-19/Israel/13075882/2020\|EPI_ISL_447442\|2020-0 | ···················· | ···················· | ···················· | ···················· | ···················· | ···················· |
| C | 3346 hCoV-19/Israel/13075879/2020\|EPI_ISL_447443\|2020-0 | ···················· | ···················· | ···················· | ···················· | ···················· | ···················· |
| C | 3347 hCoV-19/Israel/13075832/2020\|EPI_ISL_447444\|2020-0 | ···················· | ···················· | ···················· | ···················· | ···················· | ···················· |
| C | 3348 hCoV-19/Israel/13075790/2020\|EPI_ISL_447445\|2020-0 | ···················· | ···················· | ···················· | ···················· | ···················· | ···················· |
| C | 3349 hCoV-19/Israel/13075788/2020\|EPI_ISL_447446\|2020-0 | ···················· | ···················· | ···················· | ···················· | ···················· | ···················· |
| C | 3350 hCoV-19/Israel/13075782/2020\|EPI_ISL_447447\|2020-0 | ···················· | ···················· | ···················· | ···················· | ···················· | ···················· |
| C | 3351 hCoV-19/Israel/13075735/2020\|EPI_ISL_447448\|2020-0 | ···················· | ···················· | ···················· | ···················· | ···················· | ···················· |
| C | 3352 hCoV-19/Israel/13075719/2020\|EPI_ISL_447449\|2020-0 | ···················· | ···················· | ···················· | ···················· | ···················· | ······N············· |
| C | 3353 hCoV-19/Israel/13075703/2020\|EPI_ISL_447450\|2020-0 | ···················· | ···················· | ···················· | ···················· | ···················· | ···················· |
| C | 3354 hCoV-19/Israel/130710414/2020\|EPI_ISL_447460\|2020- | ···················· | ···················· | ···················· | ···················· | ···················· | ···················· |
| C | 3355 hCoV-19/Israel/130710390/2020\|EPI_ISL_447461\|2020- | ···················· | ···················· | ···················· | ···················· | ···················· | ···················· |
| C | 3356 hCoV-19/Israel/130710217/2020\|EPI_ISL_447462\|2020- | ···················· | ···················· | ···················· | ···················· | ···················· | ···················· |
| C | 3357 hCoV-19/Israel/130710211/2020\|EPI_ISL_447463\|2020- | ···················· | ···················· | ···················· | ···················· | ···················· | ···················· |

| | | | | | | | | |
|---|---|---|---|---|---|---|---|---|
| C | 3358 hCoV-19/Israel/130710159/2020\|EPI_ISL_447464\|2020- | ···················· | ···················· | ···················· | ···················· | ···················· | ···················· | |
| C | 3359 hCoV-19/Israel/130710157/2020\|EPI_ISL_447465\|2020- | ···················· | ···················· | ···················· | ···················· | ···················· | ···················· | |
| C | 3360 hCoV-19/Israel/130710097/2020\|EPI_ISL_447467\|2020- | ···················· | ···················· | ···················· | ···················· | ···················· | ···················· | |
| C | 3361 hCoV-19/Israel/130710067/2020\|EPI_ISL_447468\|2020- | ···················· | ···················· | ···················· | ···················· | ···················· | ···················· | |
| C | 3363 hCoV-19/Spain/VC-FISABIO-551/2020\|EPI_ISL_447493\|2 | ···················· | ···················· | ···················· | ···················· | ···················· | ···················· | |
| C | 3364 hCoV-19/Spain/VC-FISABIO-565/2020\|EPI_ISL_447497\|2 | ···················· | ···················· | ···················· | ···················· | ·NNN················ | ···················· | 1 |
| C | 3365 hCoV-19/Spain/VC-FISABIO-566/2020\|EPI_ISL_447498\|2 | ···················· | ···················· | ···················· | ···················· | ···················· | ···················· | |
| C | 3366 hCoV-19/England/OXON-AE532/2020\|EPI_ISL_448700\|202 | ···················· | ···················· | ···················· | ···················· | ···················· | ···················· | |
| C | 3367 hCoV-19/England/OXON-AE550/2020\|EPI_ISL_448701\|202 | ···················· | ···················· | ···················· | ···················· | ···················· | ···················· | |
| C | 3368 hCoV-19/England/OXON-AE57E/2020\|EPI_ISL_448702\|202 | ···················· | ···················· | ···················· | ···················· | ···················· | ···················· | |
| C | 3369 hCoV-19/England/OXON-AE59C/2020\|EPI_ISL_448703\|202 | ···················· | ···················· | ···················· | ···················· | ···················· | ···················· | |
| C | 3370 hCoV-19/England/OXON-AE5BA/2020\|EPI_ISL_448704\|202 | ···················· | ···················· | ···················· | ···················· | ···················· | ···················· | |
| C | 3371 hCoV-19/England/OXON-AE5D8/2020\|EPI_ISL_448705\|202 | ···················· | ···················· | ···················· | ···················· | ···················· | ···················· | |
| C | 3372 hCoV-19/England/OXON-AE5F6/2020\|EPI_ISL_448706\|202 | ···················· | ···················· | ···················· | ···················· | ···················· | ···················· | |
| C | 3373 hCoV-19/England/OXON-AE611/2020\|EPI_ISL_448707\|202 | ···················· | ···················· | ···················· | ···················· | ···················· | ···················· | |
| C | 3374 hCoV-19/England/OXON-AE65D/2020\|EPI_ISL_448708\|202 | ···················· | ···················· | ···················· | ···················· | ···················· | ···················· | |
| C | 3375 hCoV-19/England/OXON-AE67B/2020\|EPI_ISL_448709\|202 | ···················· | ···················· | ···················· | ···················· | ···················· | ···················· | |
| C | 3376 hCoV-19/England/OXON-AE699/2020\|EPI_ISL_448710\|202 | ···················· | ···················· | ···················· | ···················· | ···················· | ···················· | |
| C | 3377 hCoV-19/England/OXON-AE6D5/2020\|EPI_ISL_448711\|202 | ···················· | ···················· | ···················· | ···················· | ···················· | ···················· | |
| C | 3378 hCoV-19/England/OXON-AE6E4/2020\|EPI_ISL_448712\|202 | ···················· | ···················· | ···················· | ···················· | ···················· | ···················· | |
| C | 3379 hCoV-19/England/OXON-AE6F3/2020\|EPI_ISL_448713\|202 | ···················· | ···················· | ···················· | ···················· | ···················· | ···················· | |
| C | 3380 hCoV-19/England/OXON-AE71E/2020\|EPI_ISL_448714\|202 | ···················· | ···················· | ···················· | ···················· | ···················· | ···················· | |
| C | 3381 hCoV-19/England/OXON-AE73C/2020\|EPI_ISL_448715\|202 | ···················· | ···················· | ···················· | ···················· | ···················· | ···················· | |
| C | 3382 hCoV-19/England/OXON-AE75A/2020\|EPI_ISL_448716\|202 | ···················· | ···················· | ···················· | ···················· | ···················· | ···················· | |
| C | 3383 hCoV-19/England/OXON-AE778/2020\|EPI_ISL_448717\|202 | ···················· | ···················· | ···················· | ···················· | ···················· | ···················· | |
| C | 3384 hCoV-19/England/OXON-AE796/2020\|EPI_ISL_448718\|202 | ···················· | ···················· | ···················· | ···················· | ···················· | ···················· | |
| C | 3385 hCoV-19/England/OXON-AE7B4/2020\|EPI_ISL_448719\|202 | ···················· | ···················· | ···················· | ···················· | ···················· | ···················· | |
| C | 3386 hCoV-19/England/OXON-AE7D2/2020\|EPI_ISL_448720\|202 | ···················· | ···················· | ···················· | ···················· | ···················· | ···················· | |
| C | 3387 hCoV-19/England/OXON-AE7F0/2020\|EPI_ISL_448722\|202 | ···················· | ···················· | ···················· | ···················· | ···················· | ···················· | |
| C | 3388 hCoV-19/England/OXON-AE80C/2020\|EPI_ISL_448723\|202 | ···················· | ···················· | ···················· | ···················· | ···················· | ···················· | |
| C | 3389 hCoV-19/England/OXON-AE81B/2020\|EPI_ISL_448724\|202 | ···················· | ···················· | ···················· | ···················· | ···················· | ···················· | |
| C | 3390 hCoV-19/England/OXON-AE839/2020\|EPI_ISL_448725\|202 | ···················· | ···················· | ···················· | ···················· | ···················· | ···················· | |
| C | 3391 hCoV-19/England/OXON-AE848/2020\|EPI_ISL_448726\|202 | ···················· | ···················· | ···················· | ···················· | ···················· | ···················· | |
| C | 3392 hCoV-19/England/OXON-AE857/2020\|EPI_ISL_448727\|202 | ···················· | ···················· | ···················· | ···················· | ···················· | ···················· | |
| C | 3393 hCoV-19/England/OXON-AE875/2020\|EPI_ISL_448728\|202 | ···················· | ···················· | ···················· | ···················· | ···················· | ···················· | |
| C | 3394 hCoV-19/England/OXON-AE893/2020\|EPI_ISL_448729\|202 | ···················· | ···················· | ···················· | ···················· | ···················· | ···················· | |
| C | 3395 hCoV-19/England/OXON-AE8B1/2020\|EPI_ISL_448730\|202 | ···················· | ···················· | ···················· | ···················· | ···················· | ···················· | |
| C | 3396 hCoV-19/England/OXON-AE8DF/2020\|EPI_ISL_448731\|202 | ···················· | ···················· | ···················· | ···················· | ···················· | ···················· | |
| C | 3397 hCoV-19/England/OXON-AE8EE/2020\|EPI_ISL_448732\|202 | ···················· | ···················· | ···················· | ···················· | ···················· | ···················· | |
| C | 3398 hCoV-19/England/OXON-AE909/2020\|EPI_ISL_448733\|202 | ···················· | ···················· | ···················· | ···················· | ···················· | ···················· | |
| C | 3399 hCoV-19/England/OXON-AE927/2020\|EPI_ISL_448734\|202 | ···················· | ···················· | ···················· | ···················· | ···················· | ···················· | |
| C | 3400 hCoV-19/England/OXON-AE936/2020\|EPI_ISL_448735\|202 | ···················· | ···················· | ···················· | ···················· | ···················· | ···················· | |
| C | 3401 hCoV-19/England/OXON-AEB21/2020\|EPI_ISL_448758\|202 | ···················· | ···················· | ···················· | ···················· | ···················· | ···················· | |
| C | 3402 hCoV-19/England/OXON-AEB30/2020\|EPI_ISL_448759\|202 | ···················· | ···················· | ···················· | ···················· | ···················· | ···················· | |
| C | 3403 hCoV-19/England/OXON-AEB4F/2020\|EPI_ISL_448760\|202 | ···················· | ···················· | ···················· | ···················· | ···················· | ···················· | |
| C | 3404 hCoV-19/England/OXON-AEB7C/2020\|EPI_ISL_448762\|202 | ···················· | ···················· | ···················· | ···················· | ···················· | ···················· | |
| C | 3405 hCoV-19/England/OXON-AEB9A/2020\|EPI_ISL_448763\|202 | ···················· | ···················· | ···················· | ···················· | ···················· | ···················· | |
| C | 3406 hCoV-19/England/OXON-AEBA9/2020\|EPI_ISL_448764\|202 | ···················· | ···················· | ···················· | ···················· | ···················· | ···················· | |
| C | 3407 hCoV-19/England/OXON-AEBB8/2020\|EPI_ISL_448765\|202 | ···················· | ···················· | ···················· | ···················· | ···················· | ···················· | |
| C | 3408 hCoV-19/England/OXON-AEBC7/2020\|EPI_ISL_448766\|202 | ···················· | ···················· | ···················· | ···················· | ···················· | ···················· | |
| C | 3409 hCoV-19/England/OXON-AEBD6/2020\|EPI_ISL_448767\|202 | ···················· | ···················· | ···················· | ···················· | ···················· | ···················· | |
| C | 3410 hCoV-19/England/OXON-AEBE5/2020\|EPI_ISL_448768\|202 | ···················· | ···················· | ···················· | ···················· | ···················· | ···················· | |
| C | 3411 hCoV-19/England/OXON-AEC00/2020\|EPI_ISL_448770\|202 | ···················· | ···················· | ···················· | ···················· | ···················· | ···················· | |
| C | 3412 hCoV-19/England/OXON-AEC1F/2020\|EPI_ISL_448771\|202 | ···················· | ···················· | ···················· | ···················· | ···················· | ···················· | |
| C | 3413 hCoV-19/England/OXON-AEC3D/2020\|EPI_ISL_448772\|202 | ···················· | ···················· | ···················· | ···················· | ···················· | ···················· | |
| C | 3414 hCoV-19/England/OXON-AEC5B/2020\|EPI_ISL_448773\|202 | ···················· | ···················· | ···················· | ···················· | ···················· | ···················· | |
| C | 3415 hCoV-19/England/OXON-AEC79/2020\|EPI_ISL_448774\|202 | ···················· | ···················· | ···················· | ···················· | ···················· | ···················· | |
| C | 3416 hCoV-19/England/OXON-AEC88/2020\|EPI_ISL_448775\|202 | ···················· | ···················· | ···················· | ···················· | ···················· | ···················· | |
| C | 3417 hCoV-19/England/OXON-AEC97/2020\|EPI_ISL_448776\|202 | ···················· | ···················· | ···················· | ···················· | ···················· | ···················· | |
| C | 3418 hCoV-19/England/OXON-AECB5/2020\|EPI_ISL_448777\|202 | ···················· | ···················· | ···················· | ···················· | ···················· | ···················· | |
| C | 3419 hCoV-19/England/OXON-AECD3/2020\|EPI_ISL_448778\|202 | ···················· | ···················· | ···················· | ···················· | ···················· | ···················· | |
| C | 3420 hCoV-19/England/OXON-AECE2/2020\|EPI_ISL_448779\|202 | ···················· | ···················· | ···················· | ···················· | ···················· | ···················· | |
| C | 3421 hCoV-19/England/OXON-AED1C/2020\|EPI_ISL_448781\|202 | ···················· | ···················· | ···················· | ···················· | ···················· | ···················· | |
| C | 3422 hCoV-19/England/OXON-AED49/2020\|EPI_ISL_448782\|202 | ···················· | ···················· | ···················· | ···················· | ···················· | ···················· | |
| C | 3423 hCoV-19/England/OXON-AEE82/2020\|EPI_ISL_448790\|202 | ···················· | ···················· | ···················· | ···················· | ···················· | ···················· | |
| C | 3424 hCoV-19/England/OXON-AEE91/2020\|EPI_ISL_448791\|202 | ···················· | ···················· | ···················· | ···················· | ···················· | ···················· | |
| C | 3425 hCoV-19/England/OXON-AEEA0/2020\|EPI_ISL_448792\|202 | ···················· | ···················· | ···················· | ···················· | ···················· | ···················· | |
| C | 3426 hCoV-19/England/OXON-AEEDD/2020\|EPI_ISL_448793\|202 | ···················· | ···················· | ···················· | ···················· | ···················· | ···················· | |
| C | 3427 hCoV-19/England/OXON-AEEEC/2020\|EPI_ISL_448794\|202 | ···················· | ···················· | ···················· | ···················· | ···················· | ···················· | |
| C | 3428 hCoV-19/England/OXON-AEF07/2020\|EPI_ISL_448795\|202 | ···················· | ···················· | ···················· | ···················· | ···················· | ···················· | |
| C | 3429 hCoV-19/England/OXON-AEF34/2020\|EPI_ISL_448796\|202 | ···················· | ···················· | ···················· | ···················· | ···················· | ···················· | |
| C | 3430 hCoV-19/England/OXON-AEF43/2020\|EPI_ISL_448797\|202 | ···················· | ···················· | ···················· | ···················· | ···················· | ···················· | |
| C | 3431 hCoV-19/England/OXON-AEF8F/2020\|EPI_ISL_448799\|202 | ···················· | ···················· | ···················· | ···················· | ···················· | ···················· | |
| C | 3432 hCoV-19/India/GJ-GBRC141a/2020\|EPI_ISL_458100\|2020 | ···················· | ·A·················· | ···················· | ···················· | ···················· | ··A················· | |
| C | 3433 hCoV-19/India/GJ-GBRC141b/2020\|EPI_ISL_458101\|2020 | ···················· | ·A·················· | ···················· | ···················· | ···················· | ···················· | |
| C | 3434 hCoV-19/India/GJ-GBRC142/2020\|EPI_ISL_458102\|2020- | ···················· | ···················· | ···················· | ···················· | ···················· | ···················· | |
| C | 3435 hCoV-19/India/GJ-GBRC143/2020\|EPI_ISL_458103\|2020- | ···················· | ·A·················· | ···················· | ···················· | ···················· | ··A················· | |
| C | 3436 hCoV-19/India/GJ-GBRC144/2020\|EPI_ISL_458104\|2020- | ···················· | ···················· | ···················· | ···················· | ···················· | ···················· | |

| | | | | | | | |
|---|---|---|---|---|---|---|---|
| C | 3437 hCoV-19/India/GJ-GBRC145/2020\|EPI_ISL_458105\|2020- | ···················· | ···················· | ···················· | ···················· | ···················· | ···················· |
| C | 3438 hCoV-19/India/GJ-GBRC146a/2020\|EPI_ISL_458106\|2020 | ···················· | ···················· | ···················· | ···················· | ···················· | ···················· |
| C | 3439 hCoV-19/India/GJ-GBRC146b/2020\|EPI_ISL_458107\|2020 | ···················· | ···················· | ···················· | ···················· | ···················· | ···················· |
| C | 3440 hCoV-19/India/GJ-GBRC147/2020\|EPI_ISL_458108\|2020- | ···················· | ···················· | ···················· | ···················· | ···················· | ···················· |
| C | 3441 hCoV-19/India/GJ-GBRC148b/2020\|EPI_ISL_458110\|2020 | ···················· | ···················· | ···················· | ···················· | ···················· | ···················· |
| C | 3442 hCoV-19/India/GJ-GBRC149/2020\|EPI_ISL_458111\|2020- | ···················· | ·A·················· | ···················· | ···················· | ···················· | ···················· |
| C | 3443 hCoV-19/India/GJ-GBRC150a/2020\|EPI_ISL_458112\|2020 | ···················· | ·A·················· | ···················· | ···················· | ···················· | ··A················· |
| C | 3444 hCoV-19/India/GJ-GBRC150b/2020\|EPI_ISL_458113\|2020 | ···················· | ·A·················· | ···················· | ···················· | ···················· | ··A················· |
| C | 3445 hCoV-19/Oman/205033013/2020\|EPI_ISL_458116\|2020-05 | ···················· | ···················· | ···················· | ···················· | ···················· | ···················· |
| C | 3446 hCoV-19/Oman/205034603/2020\|EPI_ISL_458117\|2020-05 | ···················· | ···················· | ···················· | ···················· | ···················· | ···················· |
| C | 3447 hCoV-19/Oman/205033010/2020\|EPI_ISL_458118\|2020-05 | ···················· | ···················· | ···················· | ···················· | ···················· | ···················· |
| C | 3448 hCoV-19/Brazil/PA-IEC-162802/2020\|EPI_ISL_458140\|2 | ···················· | ···················· | ···················· | ···················· | ···················· | ···················· |
| C | 3449 hCoV-19/Brazil/PA-IEC-164239/2020\|EPI_ISL_458141\|2 | ···················· | ···················· | ···················· | ···················· | ···················· | ···················· |
| C | 3450 hCoV-19/Brazil/AP-IEC-162966/2020\|EPI_ISL_458142\|2 | ···················· | ···················· | ···················· | ···················· | ···················· | ···················· |
| C | 3451 hCoV-19/Brazil/AP-IEC-164082/2020\|EPI_ISL_458143\|2 | ···················· | ···················· | ···················· | ···················· | ···················· | ···················· |
| C | 3452 hCoV-19/Brazil/AP-IEC-163972/2020\|EPI_ISL_458144\|2 | ···················· | ···················· | ···················· | ···················· | ···················· | ···················· |
| C | 3453 hCoV-19/Brazil/AP-IEC-164346/2020\|EPI_ISL_458145\|2 | ···················· | ···················· | ···················· | ···················· | ···················· | ···················· |
| C | 3454 hCoV-19/Brazil/PA-IEC-164173/2020\|EPI_ISL_458146\|2 | ···················· | ···················· | ···················· | ···················· | ···················· | ···················· |
| C | 3455 hCoV-19/Brazil/PA-IEC-164218/2020\|EPI_ISL_458147\|2 | ···················· | ···················· | ···················· | ···················· | ···················· | ···················· |
| C | 3456 hCoV-19/Brazil/PA-IEC-164684/2020\|EPI_ISL_458148\|2 | ···················· | ···················· | ···················· | ···················· | ···················· | ···················· |
| C | 3457 hCoV-19/Brazil/MA-IEC-163069/2020\|EPI_ISL_458149\|2 | ···················· | ···················· | ···················· | ···················· | ···················· | ···················· |
| C | 3458 hCoV-19/Morocco/15N/2020\|EPI_ISL_458150\|2020-05-15 | ···················· | ···················· | ···················· | ···················· | ···················· | ···················· |
| C | 3459 hCoV-19/Belgium/Rega-0328186/2020\|EPI_ISL_458156\|2 | ···················· | ···················· | ···················· | ···················· | ···················· | ···················· |
| C | 3460 hCoV-19/Belgium/Rega-0328187/2020\|EPI_ISL_458157\|2 | ···················· | ···················· | ···················· | ···················· | ···················· | ···················· |
| C | 3461 hCoV-19/Belgium/Rega-0328188/2020\|EPI_ISL_458158\|2 | ···················· | ···················· | ···················· | ···················· | ···················· | ···················· |
| C | 3462 hCoV-19/Belgium/Rega-0328189/2020\|EPI_ISL_458159\|2 | ···················· | ···················· | ···················· | ···················· | ···················· | ···················· |
| C | 3463 hCoV-19/Belgium/Rega-0328190/2020\|EPI_ISL_458160\|2 | ···················· | ···················· | ···················· | ···················· | ···················· | ···················· |
| C | 3464 hCoV-19/Belgium/Rega-0328191/2020\|EPI_ISL_458161\|2 | ···················· | ···················· | ···················· | ···················· | ···················· | ···················· |
| C | 3465 hCoV-19/Belgium/Rega-0328192/2020\|EPI_ISL_458162\|2 | ···················· | ···················· | ···················· | ···················· | ···················· | ···················· |
| C | 3466 hCoV-19/Belgium/Rega-0328193/2020\|EPI_ISL_458163\|2 | ···················· | ···················· | ···················· | ···················· | ···················· | ···················· |
| C | 3467 hCoV-19/Belgium/Rega-0328194/2020\|EPI_ISL_458164\|2 | ···················· | ···················· | ···················· | ···················· | ···················· | ···················· |
| C | 3468 hCoV-19/Belgium/Rega-0328195/2020\|EPI_ISL_458165\|2 | ···················· | ···················· | ···················· | ···················· | ···················· | ···················· |
| C | 3469 hCoV-19/Belgium/Rega-0328196/2020\|EPI_ISL_458166\|2 | ···················· | ···················· | ···················· | ···················· | ···················· | ···················· |
| C | 3470 hCoV-19/Belgium/Rega-0328197/2020\|EPI_ISL_458167\|2 | ···················· | ···················· | ···················· | ···················· | ···················· | ···················· |
| C | 3471 hCoV-19/Belgium/Rega-0328198/2020\|EPI_ISL_458168\|2 | ···················· | ···················· | ···················· | ···················· | ···················· | ···················· |
| C | 3472 hCoV-19/Belgium/Rega-0328199/2020\|EPI_ISL_458169\|2 | ···················· | ···················· | ···················· | ···················· | ···················· | ···················· |
| C | 3473 hCoV-19/Belgium/Rega-0328200/2020\|EPI_ISL_458170\|2 | ···················· | ···················· | ···················· | ···················· | ···················· | ···················· |
| C | 3474 hCoV-19/Belgium/Rega-0328201/2020\|EPI_ISL_458171\|2 | ···················· | ···················· | ···················· | ···················· | ···················· | ···················· |
| C | 3475 hCoV-19/Belgium/Rega-0328202/2020\|EPI_ISL_458172\|2 | ···················· | ···················· | ···················· | ···················· | ···················· | ···················· |
| C | 3476 hCoV-19/Belgium/Rega-0328203/2020\|EPI_ISL_458173\|2 | ···················· | ···················· | ···················· | ···················· | ···················· | ···················· |
| C | 3477 hCoV-19/Belgium/Rega-0328204/2020\|EPI_ISL_458174\|2 | ···················· | ···················· | ···················· | ···················· | ···················· | ···················· |
| C | 3478 hCoV-19/Belgium/Rega-0328205/2020\|EPI_ISL_458175\|2 | ···················· | ···················· | ···················· | ···················· | ···················· | ···················· |
| C | 3479 hCoV-19/Belgium/Rega-0328206/2020\|EPI_ISL_458176\|2 | ···················· | ···················· | ···················· | ···················· | ···················· | ···················· |
| C | 3480 hCoV-19/Belgium/Rega-0328207/2020\|EPI_ISL_458177\|2 | ···················· | ···················· | ···················· | ···················· | ···················· | ···················· |
| C | 3481 hCoV-19/Belgium/Rega-0328208/2020\|EPI_ISL_458178\|2 | ···················· | ···················· | ···················· | ···················· | ···················· | ···················· |
| C | 3482 hCoV-19/Belgium/Rega-0328209/2020\|EPI_ISL_458179\|2 | ···················· | ···················· | ···················· | ···················· | ···················· | ···················· |
| C | 3483 hCoV-19/Northern Ireland/NIRE-FCC30/2020\|EPI_ISL_4 | ···················· | ···················· | ···················· | ···················· | ···················· | ···················· |
| C | 3484 hCoV-19/Northern Ireland/NIRE-FC966/2020\|EPI_ISL_4 | ···················· | ···················· | ···················· | ···················· | ···················· | ···················· |
| C | 3485 hCoV-19/England/CAMB-1ADE19/2020\|EPI_ISL_459412\|20 | ···················· | ···················· | ···················· | ···················· | ···················· | ···················· |
| C | 3486 hCoV-19/England/CAMB-1ADF07/2020\|EPI_ISL_459413\|20 | ···················· | ···················· | ···················· | ···················· | ···················· | ···················· |
| C | 3487 hCoV-19/England/CAMB-1AE434/2020\|EPI_ISL_459416\|20 | ···················· | ···················· | ···················· | ···················· | ···················· | ···················· |
| C | 3488 hCoV-19/England/CAMB-1ADE73/2020\|EPI_ISL_459421\|20 | ···················· | ···················· | ···················· | ···················· | ···················· | ···················· |
| C | 3489 hCoV-19/England/CAMB-1ADF16/2020\|EPI_ISL_459423\|20 | ···················· | ···················· | ···················· | ···················· | ···················· | ···················· |
| C | 3490 hCoV-19/England/CAMB-1AE2B2/2020\|EPI_ISL_459424\|20 | ···················· | ···················· | ···················· | ···················· | ···················· | ···················· |
| C | 3491 hCoV-19/England/CAMB-1ADE28/2020\|EPI_ISL_459425\|20 | ···················· | ···················· | ···················· | ···················· | ···················· | ···················· |
| C | 3492 hCoV-19/England/CAMB-1ADEDD/2020\|EPI_ISL_459434\|20 | ···················· | ···················· | ···················· | ···················· | ···················· | ···················· |
| C | 3493 hCoV-19/England/CAMB-1ADE37/2020\|EPI_ISL_459435\|20 | ···················· | ···················· | ···················· | ···················· | ···················· | ···················· |
| C | 3494 hCoV-19/England/CAMB-1AE030/2020\|EPI_ISL_459437\|20 | ···················· | ···················· | ···················· | ···················· | ···················· | ···················· |
| C | 3495 hCoV-19/England/CAMB-1AE540/2020\|EPI_ISL_459442\|20 | ···················· | ···················· | ···················· | ···················· | ···················· | ···················· |
| C | 3496 hCoV-19/England/CAMB-1ADDB2/2020\|EPI_ISL_459448\|20 | ···················· | ···················· | ···················· | ···················· | ···················· | ···················· |
| C | 3497 hCoV-19/England/CAMB-1AE65C/2020\|EPI_ISL_459449\|20 | ···················· | ···················· | ···················· | ···················· | ···················· | ···················· |
| C | 3498 hCoV-19/England/CAMB-1AE4CB/2020\|EPI_ISL_459476\|20 | ···················· | ···················· | ···················· | ···················· | ···················· | ···················· |
| C | 3499 hCoV-19/England/CAMB-1ADE82/2020\|EPI_ISL_459477\|20 | ···················· | ···················· | ···················· | ···················· | ···················· | ···················· |
| C | 3500 hCoV-19/England/CAMB-1ADDC1/2020\|EPI_ISL_459478\|20 | ···················· | ···················· | ···················· | ···················· | ···················· | ···················· |
| C | 3501 hCoV-19/England/CAMB-1AE003/2020\|EPI_ISL_459479\|20 | ···················· | ···················· | ···················· | ···················· | ···················· | ···················· |
| C | 3502 hCoV-19/England/CAMB-1AEA7E/2020\|EPI_ISL_459481\|20 | ···················· | ···················· | ···················· | ···················· | ···················· | ···················· |
| C | 3503 hCoV-19/England/CAMB-1AE1D3/2020\|EPI_ISL_459493\|20 | ···················· | ···················· | ···················· | ···················· | ···················· | ···················· |
| C | 3504 hCoV-19/England/CAMB-1ADE55/2020\|EPI_ISL_459496\|20 | ···················· | ···················· | ···················· | ···················· | ···················· | ···················· |
| C | 3505 hCoV-19/England/CAMB-1AE6B6/2020\|EPI_ISL_459499\|20 | ···················· | ···················· | ···················· | ···················· | ···················· | ···················· |
| C | 3506 hCoV-19/USA/MA-MGH-00497/2020\|EPI_ISL_460400\|2020- | ···················· | ···················· | ···················· | ···················· | ···················· | ···················· |
| C | 3507 hCoV-19/USA/MA-MGH-00658/2020\|EPI_ISL_460402\|2020- | ···················· | ···················· | ···················· | ···················· | ···················· | ···················· |
| C | 3508 hCoV-19/USA/MA-MGH-00694/2020\|EPI_ISL_460403\|2020- | ···················· | ···················· | ···················· | ···················· | ···················· | ···················· |
| C | 3509 hCoV-19/USA/MA-MGH-00257/2020\|EPI_ISL_460404\|2020- | ···················· | ···················· | ···················· | ···················· | ···················· | ···················· |
| C | 3510 hCoV-19/USA/MA-MGH-00684/2020\|EPI_ISL_460406\|2020- | ···················· | ···················· | ···················· | ···················· | ···················· | ···················· |
| C | 3511 hCoV-19/USA/MA-MGH-00244/2020\|EPI_ISL_460407\|2020- | ···················· | ···················· | ···················· | ···················· | ···················· | ···················· |
| C | 3512 hCoV-19/USA/MA-MGH-00282/2020\|EPI_ISL_460408\|2020- | ···················· | ···················· | ···················· | ···················· | ···················· | ···················· |
| C | 3513 hCoV-19/USA/MA-MGH-00149/2020\|EPI_ISL_460409\|2020- | ···················· | ···················· | ···················· | ···················· | ···················· | ···················· |
| C | 3514 hCoV-19/USA/MA-MGH-00200/2020\|EPI_ISL_460410\|2020- | ···················· | ···················· | ···················· | ···················· | ···················· | ···················· |

| | | | | | | | | |
|---|---|---|---|---|---|---|---|---|
| C | 3515 hCoV-19/USA/MA-MGH-00637/2020\|EPI_ISL_460412\|2020- | ···················· | ···················· | ···················· | ···················· | ···················· | ···················· | |
| C | 3516 hCoV-19/USA/MA-MGH-00533/2020\|EPI_ISL_460413\|2020- | ···················· | ···················· | ···················· | ···················· | ···················· | ···················· | |
| C | 3517 hCoV-19/USA/MA-MGH-00689/2020\|EPI_ISL_460414\|2020- | ···················· | ···················· | ···················· | ···················· | ···················· | ···················· | |
| C | 3518 hCoV-19/USA/MA-MGH-00517/2020\|EPI_ISL_460415\|2020- | ···················· | ···················· | ···················· | ···················· | ···················· | ···················· | |
| C | 3519 hCoV-19/USA/MA-MGH-00164/2020\|EPI_ISL_460417\|2020- | ···················· | ···················· | ···················· | ···················· | ···················· | ···················· | |
| C | 3520 hCoV-19/USA/MA-MGH-00316/2020\|EPI_ISL_460418\|2020- | ···················· | ···················· | ···················· | ···················· | ···················· | ···················· | |
| C | 3521 hCoV-19/USA/MA-MGH-00646/2020\|EPI_ISL_460419\|2020- | ···················· | ···················· | ···················· | ···················· | ···················· | ···················· | |
| C | 3522 hCoV-19/USA/MA-MGH-00476/2020\|EPI_ISL_460420\|2020- | ···················· | ···················· | ···················· | ···················· | ···················· | ···················· | |
| C | 3523 hCoV-19/USA/MA-MGH-00036/2020\|EPI_ISL_460421\|2020- | ···················· | ···················· | ···················· | ···················· | ···················· | ···················· | |
| C | 3524 hCoV-19/USA/MA-MGH-00696/2020\|EPI_ISL_460422\|2020- | ···················· | ···················· | ···················· | ···················· | ···················· | ···················· | |
| C | 3525 hCoV-19/USA/MA-MGH-00211/2020\|EPI_ISL_460425\|2020- | ···················· | ···················· | ···················· | ···················· | ···················· | ···················· | |
| C | 3526 hCoV-19/USA/MA-MGH-00581/2020\|EPI_ISL_460426\|2020- | ···················· | ···················· | ···················· | ···················· | ···················· | ···················· | |
| C | 3527 hCoV-19/USA/MA-MGH-00495/2020\|EPI_ISL_460427\|2020- | ···················· | ···················· | ···················· | ···················· | ···················· | ···················· | |
| C | 3528 hCoV-19/USA/MA-MGH-00441/2020\|EPI_ISL_460428\|2020- | ···················· | ···················· | ···················· | ···················· | ···················· | ···················· | |
| C | 3529 hCoV-19/USA/MA-MGH-00252/2020\|EPI_ISL_460429\|2020- | ···················· | ···················· | ···················· | ···················· | ···················· | ···················· | |
| C | 3530 hCoV-19/USA/MA-MGH-00179/2020\|EPI_ISL_460451\|2020- | ···················· | ···················· | ···················· | ···················· | ···················· | ···················· | |
| C | 3531 hCoV-19/USA/MA-MGH-00165/2020\|EPI_ISL_460453\|2020- | ···················· | ···················· | ···················· | ···················· | ···················· | ···················· | |
| C | 3532 hCoV-19/USA/MA-MGH-00459/2020\|EPI_ISL_460454\|2020- | ···················· | ···················· | ···················· | ···················· | ···················· | ···················· | |
| C | 3533 hCoV-19/USA/MA-MGH-00250/2020\|EPI_ISL_460455\|2020- | ···················· | ···················· | ···················· | ···················· | ···················· | ···················· | |
| C | 3534 hCoV-19/USA/MA-MGH-00552/2020\|EPI_ISL_460456\|2020- | ···················· | ···················· | ···················· | ···················· | ···················· | ···················· | |
| C | 3535 hCoV-19/USA/MA-MGH-00181/2020\|EPI_ISL_460457\|2020- | ···················· | ···················· | ···················· | ···················· | ···················· | ···················· | |
| C | 3536 hCoV-19/USA/MA-MGH-00525/2020\|EPI_ISL_460458\|2020- | ···················· | ···················· | ···················· | ···················· | ···················· | ···················· | |
| C | 3537 hCoV-19/USA/MA-MGH-00499/2020\|EPI_ISL_460459\|2020- | ···················· | ···················· | ···················· | ···················· | ···················· | ···················· | |
| C | 3538 hCoV-19/USA/MA-MGH-00537/2020\|EPI_ISL_460460\|2020- | ···················· | ···················· | ···················· | ···················· | ···················· | ···················· | |
| C | 3539 hCoV-19/USA/MA-MGH-00295/2020\|EPI_ISL_460461\|2020- | ···················· | ···················· | ···················· | ···················· | ···················· | ···················· | |
| C | 3540 hCoV-19/USA/MA-MGH-00150/2020\|EPI_ISL_460462\|2020- | ···················· | ···················· | ···················· | ···················· | ···················· | ···················· | |
| C | 3541 hCoV-19/USA/MA-MGH-00272/2020\|EPI_ISL_460463\|2020- | ···················· | ···················· | ···················· | ···················· | ···················· | ···················· | |
| C | 3542 hCoV-19/USA/MA-MGH-00271/2020\|EPI_ISL_460464\|2020- | ···················· | ···················· | ···················· | ···················· | ···················· | ···················· | |
| C | 3543 hCoV-19/USA/MA-MGH-00679/2020\|EPI_ISL_460465\|2020- | ···················· | ···A················ | ···················· | ···················· | ···················· | ···················· | |
| C | 3544 hCoV-19/USA/MA-MGH-00178/2020\|EPI_ISL_460466\|2020- | ···················· | ···················· | ···················· | ···················· | ···················· | ···················· | |
| C | 3545 hCoV-19/USA/MA-MGH-00004/2020\|EPI_ISL_460467\|2020- | ···················· | ···················· | ···················· | ···················· | ···················· | ···················· | |
| C | 3546 hCoV-19/USA/MA-MGH-00199/2020\|EPI_ISL_460468\|2020- | ···················· | ···················· | ···················· | ···················· | ···················· | ···················· | |
| C | 3547 hCoV-19/USA/MA-MGH-00028/2020\|EPI_ISL_460469\|2020- | ···················· | ···················· | ···················· | ···················· | ···················· | ···················· | |
| C | 3548 hCoV-19/USA/MA-MGH-00132/2020\|EPI_ISL_460470\|2020- | ···················· | ···················· | ···················· | ···················· | ···················· | ···················· | |
| C | 3549 hCoV-19/USA/MA-MGH-00205/2020\|EPI_ISL_460471\|2020- | ···················· | ···················· | ···················· | ···················· | ···················· | ···················· | |
| C | 3550 hCoV-19/USA/MA-MGH-00005/2020\|EPI_ISL_460472\|2020- | ···················· | ···················· | ···················· | ···················· | ···················· | ···················· | |
| C | 3551 hCoV-19/Scotland/EDB4546/2020\|EPI_ISL_461720\|2020- | ···················· | ···················· | ···················· | ···················· | ···················· | ···················· | |
| C | 3552 hCoV-19/Scotland/EDB4547/2020\|EPI_ISL_461721\|2020- | ···················· | ···················· | ···················· | ···················· | ···················· | ···················· | |
| C | 3553 hCoV-19/Scotland/EDB4552/2020\|EPI_ISL_461723\|2020- | ···················· | ·A·················· | ···················· | ···················· | ···················· | ···················· | |
| C | 3554 hCoV-19/Scotland/EDB4558/2020\|EPI_ISL_461726\|2020- | ···················· | ·A·················· | ···················· | ···················· | ··············N····· | ···················· | |
| C | 3555 hCoV-19/Scotland/EDB4560/2020\|EPI_ISL_461728\|2020- | ···················· | ···················· | ···················· | ···················· | ···················· | ···················· | |
| C | 3556 hCoV-19/Scotland/EDB4563/2020\|EPI_ISL_461729\|2020- | ···················· | ·A·················· | ···················· | ···················· | ···················· | ···················· | |
| C | 3557 hCoV-19/Scotland/EDB4565/2020\|EPI_ISL_461730\|2020- | ···················· | ·A·················· | ···················· | ···················· | ···················· | ···················· | |
| C | 3558 hCoV-19/Scotland/EDB4922/2020\|EPI_ISL_461731\|2020- | ···················· | ···················· | ···················· | ···················· | ···················· | ···················· | |
| C | 3559 hCoV-19/Scotland/EDB5381/2020\|EPI_ISL_461733\|2020- | ···················· | ···················A | ···················· | ···················· | ···················· | ···················· | 1 |
| C | 3560 hCoV-19/Scotland/EDB5388/2020\|EPI_ISL_461735\|2020- | ···················· | ···················A | ···················· | ···················· | ···················· | ···················· | 1 |
| C | 3561 hCoV-19/Scotland/EDB5389/2020\|EPI_ISL_461736\|2020- | ···················· | ···················A | ···················· | ···················· | ···················· | ···················· | 1 |
| C | 3562 hCoV-19/Scotland/EDB5390/2020\|EPI_ISL_461737\|2020- | ···················· | ···················A | ···················· | ···················· | ···················· | ···················· | 1 |
| C | 3563 hCoV-19/Scotland/EDB5391/2020\|EPI_ISL_461738\|2020- | ···················· | ···················· | ···················· | ···················· | ···················· | ···················· | |
| C | 3564 hCoV-19/Scotland/EDB5448/2020\|EPI_ISL_461739\|2020- | ···················· | ·A·················· | ···················· | ···················· | ···················· | ···················· | |
| C | 3565 hCoV-19/Scotland/EDB5462/2020\|EPI_ISL_461741\|2020- | ···················· | ···················A | ···················· | ···················· | ···················· | ···················· | 1 |
| C | 3566 hCoV-19/Scotland/EDB5464/2020\|EPI_ISL_461742\|2020- | ···················· | ···················· | ···················· | ···················· | ···················· | ···················· | |
| C | 3567 hCoV-19/Scotland/EDB5466/2020\|EPI_ISL_461743\|2020- | ···················· | ···················· | ···················· | ···················· | ···················· | ···················· | |
| C | 3568 hCoV-19/Scotland/EDB5469/2020\|EPI_ISL_461744\|2020- | ···················· | ···················· | ···················· | ···················· | ···················· | ···················· | |
| C | 3571 hCoV-19/Scotland/EDB5474/2020\|EPI_ISL_461748\|2020- | ···················· | ···················· | ···················· | ···················· | ···················· | ···················· | |
| C | 3572 hCoV-19/Scotland/EDB5475/2020\|EPI_ISL_461749\|2020- | ···················· | ·············A······ | ···················· | ···················· | ···················· | ···················· | |
| C | 3573 hCoV-19/Scotland/EDB5478/2020\|EPI_ISL_461751\|2020- | ···················· | ···················· | ···················· | ···················· | ···················· | ···················· | |
| C | 3574 hCoV-19/Scotland/EDB5481/2020\|EPI_ISL_461753\|2020- | ···················· | ···················· | ···················· | ···················· | ···················· | ···················· | |
| C | 3575 hCoV-19/Scotland/EDB5482/2020\|EPI_ISL_461754\|2020- | ···················· | ···················A | ···················· | ···················· | ···················· | ···················· | 1 |
| C | 3576 hCoV-19/Scotland/EDB5483/2020\|EPI_ISL_461755\|2020- | ···················· | ···················· | ···················· | ···················· | ···················· | ···················· | |
| C | 3577 hCoV-19/Scotland/EDB5484/2020\|EPI_ISL_461756\|2020- | ···················· | ···················· | ···················· | ···················· | ···················· | ···················· | |
| C | 3578 hCoV-19/Scotland/EDB5486/2020\|EPI_ISL_461757\|2020- | ···················· | ···················· | ···················· | ···················· | ···················· | ···················· | |
| C | 3579 hCoV-19/Scotland/EDB5590/2020\|EPI_ISL_461759\|2020- | ···················· | ·A·················· | ···················· | ···················· | ···················· | ···················· | |
| C | 3580 hCoV-19/England/NORT-285423/2020\|EPI_ISL_461791\|20 | ···················· | ···················· | ···················· | ···················· | ···················· | ···················· | |
| C | 3581 hCoV-19/England/NORT-2854E7/2020\|EPI_ISL_461792\|20 | ···················· | ···················· | ···················· | ···················· | ···················· | ···················· | |
| C | 3582 hCoV-19/England/NORT-28597F/2020\|EPI_ISL_461793\|20 | ···················· | ···················· | ···················· | ···················· | ···················· | ···················· | |
| C | 3583 hCoV-19/England/NORT-28ADBA/2020\|EPI_ISL_461795\|20 | ···················· | ···················· | ···················· | ···················· | ···················· | ···················· | |
| C | 3584 hCoV-19/England/NORT-28AF0F/2020\|EPI_ISL_461797\|20 | ···················· | ···················· | ···················· | ···················· | ···················· | ···················· | |
| C | 3585 hCoV-19/Israel/2046129/2020\|EPI_ISL_447406\|2020-03 | ···················· | ···················· | ···················· | ···················· | ···················· | ···················· | |
| C | 3586 hCoV-19/Israel/2023920/2020\|EPI_ISL_447408\|2020-03 | ···················· | ···················· | ···················· | ···················· | ···················· | ···················· | |
| C | 3587 hCoV-19/Israel/2020087/2020\|EPI_ISL_447409\|2020-03 | ···················· | ···················· | ···················· | ···················· | ···················· | ···················· | |
| C | 3588 hCoV-19/Israel/2020084/2020\|EPI_ISL_447410\|2020-03 | ···················· | ···················· | ···················· | ···················· | ···················· | ···················· | |
| C | 3589 hCoV-19/Israel/2020078/2020\|EPI_ISL_447411\|2020-03 | ···················· | ···················· | ···················· | ···················· | ···················· | ···················· | |
| C | 3590 hCoV-19/Israel/2020069/2020\|EPI_ISL_447412\|2020-03 | ···················· | ···················· | ···················· | ···················· | ···················· | ···················· | |
| C | 3591 hCoV-19/Israel/2020038/2020\|EPI_ISL_447416\|2020-03 | ···················· | ···················· | ···················· | ···················· | ···················· | ···················· | |
| C | 3592 hCoV-19/Israel/1639996/2020\|EPI_ISL_447417\|2020-03 | ···················· | ···················· | ···················· | ···················· | ···················· | ···················· | |
| C | 3593 hCoV-19/Israel/1639953/2020\|EPI_ISL_447418\|2020-03 | ···················· | ···················· | ···················· | ···················· | ···················· | ·········N·········· | |
| C | 3594 hCoV-19/Israel/13077882/2020\|EPI_ISL_447419\|2020-0 | ···················· | ···················· | ···················· | ···················· | ···················· | ···················· | |

| | | | | | | | | |
|---|---|---|---|---|---|---|---|---|
| C | 3595 hCoV-19/Israel/13077875/2020\|EPI_ISL_447420\|2020-0 | .................... | .................... | .................... | .................... | .................... | .................... | |
| C | 3596 hCoV-19/Israel/13077846/2020\|EPI_ISL_447422\|2020-0 | .................... | .................... | .................... | .................... | .................... | .................... | |
| C | 3597 hCoV-19/Israel/13077840/2020\|EPI_ISL_447423\|2020-0 | .................... | .................... | .................... | .................... | .................... | .................... | |
| C | 3598 hCoV-19/Israel/13077823/2020\|EPI_ISL_447424\|2020-0 | .................... | .................... | .................... | .................... | .................... | .................... | |
| C | 3599 hCoV-19/Israel/13077726/2020\|EPI_ISL_447426\|2020-0 | .................... | .................... | .................... | .................... | .................... | .................... | |
| C | 3600 hCoV-19/England/OXON-AE954/2020\|EPI_ISL_448736\|202 | .................... | .................... | .................... | .................... | .................... | .................... | |
| C | 3601 hCoV-19/England/OXON-AE963/2020\|EPI_ISL_448737\|202 | .................... | .................... | .................... | .................... | .................... | .................... | |
| C | 3602 hCoV-19/England/OXON-AE972/2020\|EPI_ISL_448738\|202 | .................... | .................... | .................... | .................... | .................... | .................... | |
| C | 3603 hCoV-19/England/OXON-AE981/2020\|EPI_ISL_448739\|202 | .................... | .................... | .................... | .................... | .................... | .................... | |
| C | 3604 hCoV-19/England/OXON-AE990/2020\|EPI_ISL_448740\|202 | .................... | .................... | .................... | .................... | .................... | .................... | |
| C | 3605 hCoV-19/England/OXON-AE9CD/2020\|EPI_ISL_448743\|202 | .................... | .................... | .................... | .................... | .................... | .................... | |
| C | 3606 hCoV-19/England/OXON-AE9DC/2020\|EPI_ISL_448744\|202 | .................... | .................... | .................... | .................... | .................... | .................... | |
| C | 3607 hCoV-19/England/OXON-AE9FA/2020\|EPI_ISL_448745\|202 | .................... | .................... | .................... | .................... | .................... | .................... | |
| C | 3608 hCoV-19/England/OXON-AEA06/2020\|EPI_ISL_448746\|202 | .................... | .................... | .................... | .................... | .................... | .................... | |
| C | 3609 hCoV-19/England/OXON-AEA15/2020\|EPI_ISL_448747\|202 | .................... | .................... | .................... | .................... | .................... | .................... | |
| C | 3610 hCoV-19/England/OXON-AEA33/2020\|EPI_ISL_448749\|202 | .................... | .................... | .................... | .................... | .................... | .................... | |
| C | 3611 hCoV-19/England/OXON-AEA51/2020\|EPI_ISL_448750\|202 | .................... | .................... | .................... | .................... | .................... | .................... | |
| C | 3612 hCoV-19/England/OXON-AEA8E/2020\|EPI_ISL_448751\|202 | .................... | .................... | .................... | .................... | .................... | .................... | |
| C | 3613 hCoV-19/England/OXON-AEA9D/2020\|EPI_ISL_448752\|202 | .................... | .................... | .................... | .................... | .................... | .................... | |
| C | 3614 hCoV-19/England/OXON-AEAD9/2020\|EPI_ISL_448754\|202 | .................... | .................... | .................... | .................... | .................... | .................... | |
| C | 3615 hCoV-19/England/OXON-AEAF7/2020\|EPI_ISL_448755\|202 | .................... | .................... | .................... | .................... | .................... | .................... | |
| C | 3616 hCoV-19/England/OXON-AEB03/2020\|EPI_ISL_448756\|202 | .................... | .................... | .................... | .................... | .................... | .................... | |
| C | 3617 hCoV-19/England/OXON-AEB12/2020\|EPI_ISL_448757\|202 | .................... | .................... | .................... | .................... | .................... | .................... | |
| C | 3618 hCoV-19/England/OXON-AEBF4/2020\|EPI_ISL_448769\|202 | .................... | .................... | .................... | .................... | .................... | .................... | |
| C | 3619 hCoV-19/England/CAMB-1B1B48/2020\|EPI_ISL_453000\|20 | .................... | .................... | .................... | .................... | .................... | .................... | |
| C | 3620 hCoV-19/England/CAMB-1B2048/2020\|EPI_ISL_453001\|20 | .................... | .................... | .................... | .................... | .................... | .................... | |
| C | 3622 hCoV-19/England/CAMB-1B20DF/2020\|EPI_ISL_453004\|20 | .................... | .................... | .................... | .................... | .................... | .................... | |
| C | 3623 hCoV-19/England/CAMB-1B2109/2020\|EPI_ISL_453005\|20 | .................... | .................... | .................... | .................... | .................... | .................... | |
| C | 3624 hCoV-19/Scotland/CVR1298/2020\|EPI_ISL_453006\|2020- | .................... | .................... | .................... | .................... | .................... | .................... | |
| C | 3625 hCoV-19/Scotland/CVR1923/2020\|EPI_ISL_453013\|2020- | .................... | .................... | .................... | .................... | .................... | .................... | |
| C | 3626 hCoV-19/Scotland/CVR2055/2020\|EPI_ISL_453019\|2020- | .................... | .................... | .................... | .................... | .................... | .................... | |
| C | 3627 hCoV-19/Scotland/CVR2237/2020\|EPI_ISL_453030\|2020- | .................... | .................... | .................... | .................... | .................... | .................... | |
| C | 3628 hCoV-19/Scotland/CVR2244/2020\|EPI_ISL_453031\|2020- | .................... | ...................A | .................... | .................... | .................... | .................... | 1 |
| C | 3630 hCoV-19/Scotland/CVR2258/2020\|EPI_ISL_453035\|2020- | .................... | .................... | .................... | .................... | .................... | .................... | |
| C | 3631 hCoV-19/Scotland/CVR2263/2020\|EPI_ISL_453037\|2020- | .................... | .................... | .................... | .................... | NN.................. | NN.................. | |
| C | 3632 hCoV-19/Scotland/CVR2274/2020\|EPI_ISL_453040\|2020- | .................... | .................... | .................... | .................... | .................... | .................... | |
| C | 3633 hCoV-19/Scotland/CVR2286/2020\|EPI_ISL_453041\|2020- | .................... | .................... | .................... | .................... | .................... | .................... | |
| C | 3634 hCoV-19/Scotland/CVR2291/2020\|EPI_ISL_453042\|2020- | .................... | .................... | .................... | .................... | .................... | .................... | |
| C | 3635 hCoV-19/Scotland/CVR2304/2020\|EPI_ISL_453044\|2020- | .................... | .................... | .................... | .................... | .................... | .................... | |
| C | 3636 hCoV-19/Portugal/PT0577/2020\|EPI_ISL_454300\|2020-0 | .................... | .................... | .................... | .................... | .................... | .................... | |
| C | 3637 hCoV-19/Portugal/PT0578/2020\|EPI_ISL_454301\|2020-0 | .................... | .................... | .................... | .................... | .................... | .................... | |
| C | 3638 hCoV-19/Portugal/PT0579/2020\|EPI_ISL_454302\|2020-0 | .................... | .................... | .................... | .................... | .................... | .................... | |
| C | 3639 hCoV-19/Portugal/PT0580/2020\|EPI_ISL_454303\|2020-0 | .................... | .................... | .................... | .................... | .................... | .................... | |
| C | 3640 hCoV-19/Portugal/PT0581/2020\|EPI_ISL_454304\|2020-0 | .................... | .................... | .................... | .................... | .................... | .................... | |
| C | 3641 hCoV-19/Portugal/PT0584/2020\|EPI_ISL_454307\|2020-0 | .................... | .................... | .................... | .................... | .................... | .................... | |
| C | 3642 hCoV-19/Portugal/PT0588/2020\|EPI_ISL_454311\|2020-0 | .................... | .................... | .................... | .................... | .................... | .................... | |
| C | 3643 hCoV-19/Portugal/PT0591/2020\|EPI_ISL_454314\|2020-0 | .................... | .................... | .................... | .................... | .................... | .................... | |
| C | 3644 hCoV-19/Portugal/PT0592/2020\|EPI_ISL_454315\|2020-0 | .................... | .................... | .................... | .................... | .................... | .................... | |
| C | 3645 hCoV-19/Portugal/PT0594/2020\|EPI_ISL_454317\|2020-0 | .................... | .................... | .................... | .................... | .................... | .................... | |
| C | 3646 hCoV-19/Portugal/PT0595/2020\|EPI_ISL_454318\|2020-0 | .................... | .................... | .................... | .................... | .................... | .................... | |
| C | 3647 hCoV-19/Portugal/PT0596/2020\|EPI_ISL_454319\|2020-0 | .................... | .................... | .................... | .................... | .................... | .................... | |
| C | 3648 hCoV-19/Portugal/PT0597/2020\|EPI_ISL_454320\|2020-0 | .................... | .................... | .................... | .................... | .................... | .................... | |
| C | 3649 hCoV-19/Portugal/PT0598/2020\|EPI_ISL_454321\|2020-0 | .................... | .................... | .................... | .................... | .................... | .................... | |
| C | 3650 hCoV-19/Portugal/PT0599/2020\|EPI_ISL_454322\|2020-0 | .................... | .................... | .................... | .................... | .................... | .................... | |
| C | 3651 hCoV-19/Portugal/PT0601/2020\|EPI_ISL_454324\|2020-0 | .................... | .................... | .................... | .................... | .................... | .................... | |
| C | 3652 hCoV-19/Portugal/PT0603/2020\|EPI_ISL_454326\|2020-0 | .................... | .................... | .................... | .................... | .................... | .................... | |
| C | 3653 hCoV-19/Portugal/PT0604/2020\|EPI_ISL_454327\|2020-0 | .................... | .................... | .................... | .................... | .................... | .................... | |
| C | 3654 hCoV-19/Portugal/PT0605/2020\|EPI_ISL_454328\|2020-0 | .................... | .................... | .................... | .................... | .................... | .................... | |
| C | 3655 hCoV-19/Portugal/PT0606/2020\|EPI_ISL_454329\|2020-0 | .................... | .................... | .................... | .................... | .................... | .................... | |
| C | 3656 hCoV-19/Portugal/PT0607/2020\|EPI_ISL_454330\|2020-0 | .................... | .................... | .................... | .................... | .................... | .................... | |
| C | 3657 hCoV-19/Portugal/PT0608/2020\|EPI_ISL_454331\|2020-0 | .................... | .................... | .................... | .................... | .................... | .................... | |
| C | 3658 hCoV-19/Portugal/PT0609/2020\|EPI_ISL_454332\|2020-0 | .................... | .................... | .................... | .................... | .................... | .................... | |
| C | 3659 hCoV-19/Portugal/PT0610/2020\|EPI_ISL_454333\|2020-0 | .................... | .................... | .................... | .................... | .................... | .................... | |
| C | 3660 hCoV-19/Portugal/PT0611/2020\|EPI_ISL_454334\|2020-0 | .................... | .................... | .................... | .................... | .................... | .................... | |
| C | 3661 hCoV-19/Portugal/PT0612/2020\|EPI_ISL_454335\|2020-0 | .................... | .................... | .................... | .................... | .................... | .................... | |
| C | 3662 hCoV-19/Portugal/PT0614/2020\|EPI_ISL_454337\|2020-0 | .................... | .................... | .................... | .................... | .................... | .................... | |
| C | 3663 hCoV-19/Portugal/PT0615/2020\|EPI_ISL_454338\|2020-0 | .................... | .................... | .................... | .................... | .................... | .................... | |
| C | 3664 hCoV-19/Portugal/PT0616/2020\|EPI_ISL_454339\|2020-0 | .................... | .................... | .................... | .................... | .................... | .................... | |
| C | 3665 hCoV-19/Portugal/PT0617/2020\|EPI_ISL_454340\|2020-0 | .................... | .................... | .................... | .................... | .................... | .................... | |
| C | 3666 hCoV-19/Portugal/PT0618/2020\|EPI_ISL_454341\|2020-0 | .................... | .................... | .................... | .................... | .................... | .................... | |
| C | 3667 hCoV-19/Portugal/PT0619/2020\|EPI_ISL_454342\|2020-0 | .................... | .................... | .................... | .................... | .................... | .................... | |
| C | 3668 hCoV-19/Portugal/PT0620/2020\|EPI_ISL_454343\|2020-0 | .................... | .................... | .................... | .................... | .................... | .................... | |
| C | 3669 hCoV-19/Portugal/PT0621/2020\|EPI_ISL_454344\|2020-0 | .................... | .................... | .................... | .................... | .................... | .................... | |
| C | 3670 hCoV-19/Portugal/PT0622/2020\|EPI_ISL_454345\|2020-0 | .................... | .................... | .................... | .................... | .................... | .................... | |
| C | 3671 hCoV-19/Portugal/PT0623/2020\|EPI_ISL_454346\|2020-0 | .................... | .................... | .................... | .................... | .................... | .................... | |
| C | 3672 hCoV-19/Portugal/PT0624/2020\|EPI_ISL_454347\|2020-0 | .................... | .................... | .................... | .................... | .................... | .................... | |
| C | 3673 hCoV-19/Portugal/PT0625/2020\|EPI_ISL_454348\|2020-0 | .................... | .................... | .................... | .................... | .................... | .................... | |
| C | 3674 hCoV-19/Portugal/PT0626/2020\|EPI_ISL_454349\|2020-0 | .................... | .................... | .................... | .................... | .................... | .................... | |

| | | | | | | | | |
|---|---|---|---|---|---|---|---|---|
| C | 3675 | hCoV-19/Portugal/PT0627/2020\|EPI_ISL_454350\|2020-0 | ···················· | ···················· | ···················· | ···················· | ···················· | ···················· |
| C | 3676 | hCoV-19/Portugal/PT0628/2020\|EPI_ISL_454351\|2020-0 | ···················· | ···················· | ···················· | ···················· | ···················· | ···················· |
| C | 3677 | hCoV-19/Portugal/PT0629/2020\|EPI_ISL_454352\|2020-0 | ···················· | ···················· | ···················· | ···················· | ···················· | ···················· |
| C | 3678 | hCoV-19/USA/PA-MGEL62/2020\|EPI_ISL_454353\|2020-03- | ···················· | ···················· | ···················· | ···················· | ···················· | ···················· |
| C | 3679 | hCoV-19/USA/PA-MGEL63/2020\|EPI_ISL_454354\|2020-03- | ···················· | ···················· | ···················· | ···················· | ···················· | ···················· |
| C | 3680 | hCoV-19/USA/PA-MGEL65/2020\|EPI_ISL_454355\|2020-03- | ···················· | ···················· | ···················· | ···················· | ···················· | ···················· |
| C | 3681 | hCoV-19/USA/PA-MGEL66/2020\|EPI_ISL_454356\|2020-03- | ···················· | ···················· | ···················· | ···················· | ···················· | ···················· |
| C | 3682 | hCoV-19/USA/PA-MGEL68/2020\|EPI_ISL_454357\|2020-03- | ···················· | ···················· | ···················· | ···················· | ···················· | ···················· |
| C | 3683 | hCoV-19/USA/PA-MGEL70/2020\|EPI_ISL_454358\|2020-03- | ···················· | ···················· | ···················· | ···················· | ···················· | ···················· |
| C | 3684 | hCoV-19/USA/PA-MGEL73/2020\|EPI_ISL_454359\|2020-03- | ···················· | ···················· | ···················· | ···················· | ···················· | ···················· |
| C | 3685 | hCoV-19/USA/PA-MGEL74/2020\|EPI_ISL_454360\|2020-03- | ···················· | ···················· | ···················· | ···················· | ···················· | ···················· |
| C | 3686 | hCoV-19/USA/PA-MGEL75/2020\|EPI_ISL_454361\|2020-03- | ···················· | ···················· | ···················· | ···················· | ···················· | ···················· |
| C | 3687 | hCoV-19/USA/PA-MGEL76/2020\|EPI_ISL_454362\|2020-03- | ···················· | ···················· | ···················· | ···················· | ···················· | ···················· |
| C | 3688 | hCoV-19/USA/PA-MGEL77/2020\|EPI_ISL_454363\|2020-03- | ···················· | ···················· | ···················· | ···················· | ···················· | ···················· |
| C | 3689 | hCoV-19/USA/PA-MGEL78/2020\|EPI_ISL_454364\|2020-03- | ···················· | ···················· | ···················· | ···················· | ···················· | ···················· |
| C | 3690 | hCoV-19/USA/PA-MGEL80/2020\|EPI_ISL_454365\|2020-03- | ···················· | ···················· | ···················· | ···················· | ···················· | ···················· |
| C | 3691 | hCoV-19/USA/PA-MGEL81/2020\|EPI_ISL_454366\|2020-03- | ···················· | ···················· | ···················· | ···················· | ···················· | ···················· |
| C | 3692 | hCoV-19/USA/PA-MGEL83/2020\|EPI_ISL_454367\|2020-03- | ···················· | ···················· | ···················· | ···················· | ···················· | ···················· |
| C | 3693 | hCoV-19/USA/PA-MGEL84/2020\|EPI_ISL_454368\|2020-03- | ···················· | ···················· | ···················· | ···················· | ···················· | ···················· |
| C | 3694 | hCoV-19/USA/PA-MGEL85/2020\|EPI_ISL_454369\|2020 | ···················· | ···················· | ···················· | ···················· | ···················· | ···················· |
| C | 3695 | hCoV-19/USA/PA-MGEL86/2020\|EPI_ISL_454370\|2020-04- | ···················· | ···················· | ···················· | ···················· | ···················· | ···················· |
| C | 3696 | hCoV-19/USA/PA-MGEL88/2020\|EPI_ISL_454371\|2020-04- | ···················· | ···················· | ···················· | ···················· | ···················· | ···················· |
| C | 3697 | hCoV-19/USA/PA-MGEL89/2020\|EPI_ISL_454372\|2020-03- | ···················· | ···················· | ···················· | ···················· | ···················· | ···················· |
| C | 3698 | hCoV-19/USA/PA-MGEL91/2020\|EPI_ISL_454373\|2020-03- | ···················· | ···················· | ···················· | ···················· | ···················· | ···················· |
| C | 3699 | hCoV-19/USA/PA-MGEL92/2020\|EPI_ISL_454374\|2020-03- | ···················· | ···················· | ···················· | ···················· | ···················· | ···················· |
| C | 3700 | hCoV-19/USA/PA-MGEL94/2020\|EPI_ISL_454375\|2020-04- | ···················· | ···················· | ···················· | ···················· | ···················· | ···················· |
| C | 3701 | hCoV-19/USA/PA-MGEL96/2020\|EPI_ISL_454376\|2020-03- | ···················· | ···················· | ···················· | ···················· | ···················· | ···················· |
| C | 3702 | hCoV-19/USA/PA-MGEL97/2020\|EPI_ISL_454377\|2020 | ···················· | ···················· | ···················· | ···················· | ···················· | ···················· |
| C | 3703 | hCoV-19/USA/PA-MGEL98/2020\|EPI_ISL_454378\|2020-03- | ···················· | ···················· | ···················· | ···················· | ···················· | ···················· |
| C | 3704 | hCoV-19/USA/LA-BIE-069/2020\|EPI_ISL_455622\|2020-04 | ···················· | ···················· | ···················· | ···················· | ···················· | ···················· |
| C | 3705 | hCoV-19/USA/LA-BIE-070/2020\|EPI_ISL_455623\|2020-04 | ···················· | ···················· | ···················· | ···················· | ···················· | ···················· |
| C | 3706 | hCoV-19/Thailand/Bangkok_2295/2020\|EPI_ISL_455624\| | ···················· | ···················· | ···················· | ···················· | ···················· | ···················· |
| C | 3707 | hCoV-19/Portugal/PT0201/2020\|EPI_ISL_455625\|2020-0 | ···················· | ···················· | ···················· | ···················· | ···················· | ···················· |
| C | 3708 | hCoV-19/Portugal/PT0202/2020\|EPI_ISL_455626\|2020-0 | ···················· | ···················· | ···················· | ···················· | ···················· | ···················· |
| C | 3709 | hCoV-19/Portugal/PT0549/2020\|EPI_ISL_455627\|2020-0 | ···················· | ···················· | ···················· | ···················· | ···················· | ···················· |
| C | 3710 | hCoV-19/South Africa/KRISP-101/2020\|EPI_ISL_455631 | ···················· | ···················· | ···················· | ···················· | ···················· | ···················· |
| C | 3711 | hCoV-19/South Africa/KRISP-102/2020\|EPI_ISL_455632 | ···················· | ···················· | ···················· | ···················· | ···················· | ···················· |
| C | 3712 | hCoV-19/South Africa/KRISP-103/2020\|EPI_ISL_455633 | ···················· | ···················· | ···················· | ···················· | ···················· | ···················· |
| C | 3713 | hCoV-19/South Africa/KRISP-105/2020\|EPI_ISL_455635 | ···················· | ···················· | ···················· | ···················· | ···················· | ···················· |
| C | 3714 | hCoV-19/South Africa/KRISP-106/2020\|EPI_ISL_455636 | ···················· | ···················· | ···················· | ···················· | ···················· | ···················· |
| C | 3715 | hCoV-19/South Africa/KRISP-109/2020\|EPI_ISL_455639 | ···················· | ···················· | ···················· | ···················· | ···················· | ···················· |
| C | 3716 | hCoV-19/India/WB-S4/2020\|EPI_ISL_455640\|2020-03-27 | ···················· | ···················· | ···················· | ···················· | ···················· | ···················· |
| C | 3717 | hCoV-19/India/WB-S7/2020\|EPI_ISL_455641\|2020-03-31 | ···················· | ···················· | ···················· | ···················· | ···················· | ···················· |
| C | 3718 | hCoV-19/India/WB-S13/2020\|EPI_ISL_455643\|2020-03-3 | ···················· | ···················· | ···················· | ···················· | ···················· | ···················· |
| C | 3719 | hCoV-19/India/WB-S14/2020\|EPI_ISL_455644\|2020-04-0 | ···················· | ···················· | ···················· | ···················· | ···················· | ···················· |
| C | 3720 | hCoV-19/India/WB-S15/2020\|EPI_ISL_455645\|2020-04-0 | ···················· | ···················· | ···················· | ···················· | ···················· | ···················· |
| C | 3721 | hCoV-19/India/WB-S16/2020\|EPI_ISL_455646\|2020-04-0 | ···················· | ···················· | ···················· | ···················· | ···················· | ···················· |
| C | 3722 | hCoV-19/India/WB-S17/2020\|EPI_ISL_455647\|2020-04-1 | ···················· | ···················· | ···················· | ···················· | ···················· | ···················· |
| C | 3723 | hCoV-19/India/WB-S18/2020\|EPI_ISL_455648\|2020-04-2 | ···················· | ···················· | ···················· | ···················· | ··A················· | ···················· |
| C | 3724 | hCoV-19/India/WB-S19/2020\|EPI_ISL_455649\|2020-04-2 | ···················· | ···················· | ···················· | ···················· | ···················· | ···················· |
| C | 3725 | hCoV-19/India/WB-S20/2020\|EPI_ISL_455650\|2020-04-2 | ···················· | ···················· | ···················· | ···················· | ···················· | ···················· |
| C | 3726 | hCoV-19/India/WB-S21/2020\|EPI_ISL_455651\|2020-04-2 | ···················· | ···················· | ···················· | ···················· | ···················· | ···················· |
| C | 3727 | hCoV-19/India/WB-S22/2020\|EPI_ISL_455652\|2020-04-2 | ···················· | ···················· | ···················· | ···················· | ···················· | ···················· |
| C | 3728 | hCoV-19/India/WB-S23/2020\|EPI_ISL_455653\|2020-04-2 | ···················· | ···················· | ···················· | ···················· | ···················· | ···················· |
| C | 3729 | hCoV-19/India/WB-S24/2020\|EPI_ISL_455654\|2020-04-2 | ···················· | ···················· | ···················· | ···················· | ···················· | ···················· |
| C | 3730 | hCoV-19/India/RJ-S25/2020\|EPI_ISL_455655\|2020-04-3 | ···················· | ···················· | ···················· | ···················· | ···················· | ···················· |
| C | 3731 | hCoV-19/India/WB-S27/2020\|EPI_ISL_455656\|2020-04-3 | ···················· | ···················· | ···················· | ···················· | ···················· | ···················· |
| C | 3732 | hCoV-19/India/WB-S28/2020\|EPI_ISL_455657\|2020-04-3 | ···················· | ···················· | ···················· | ···················· | ···················· | ···················· |
| C | 3733 | hCoV-19/India/WB-S29/2020\|EPI_ISL_455658\|2020-04-3 | ···················· | ···················· | ···················· | ···················· | ···················· | ···················· |
| C | 3734 | hCoV-19/India/WB-S30/2020\|EPI_ISL_455659\|2020-05-0 | ···················· | ···················· | ···················· | ···················· | ···················· | ···················· |
| C | 3735 | hCoV-19/India/WB-S31/2020\|EPI_ISL_455660\|2020-05-0 | ···················· | ···················· | ···················· | ···················· | ···················· | ···················· |
| C | 3736 | hCoV-19/India/WB-S32/2020\|EPI_ISL_455661\|2020-05-0 | ···················· | ···················· | ···················· | ···················· | ···················· | ···················· |
| C | 3737 | hCoV-19/India/WB-S33/2020\|EPI_ISL_455662\|2020-05-0 | ···················· | ···················· | ···················· | ···················· | ···················· | ···················· |
| C | 3738 | hCoV-19/India/WB-S34/2020\|EPI_ISL_455663\|2020-05-0 | ···················· | ···················· | ···················· | ···················· | ···················· | ···················· |
| C | 3739 | hCoV-19/India/WB-S42/2020\|EPI_ISL_455664\|2020-05-0 | ···················· | ···················· | ···················· | ···················· | ···················· | ···················· |
| C | 3740 | hCoV-19/India/WB-S44/2020\|EPI_ISL_455665\|2020-05-0 | ···················· | ···················· | ···················· | ···················· | ···················· | ···················· |
| C | 3741 | hCoV-19/India/WB-S45/2020\|EPI_ISL_455666\|2020-05-0 | ···················· | ···················· | ···················· | ···················· | ··A················· | ···················· |
| C | 3742 | hCoV-19/India/WB-S46/2020\|EPI_ISL_455667\|2020-05-0 | ···················· | ···················· | ···················· | ···················· | ···················· | ···················· |
| C | 3743 | hCoV-19/India/WB-S48/2020\|EPI_ISL_455668\|2020-05-0 | ···················· | ···················· | ···················· | ···················· | ···················· | ···················· |
| C | 3744 | hCoV-19/India/WB-S49/2020\|EPI_ISL_455669\|2020-05-0 | ···················· | ···················· | ···················· | ···················· | ··A················· | ···················· |
| C | 3745 | hCoV-19/India/WB-S50/2020\|EPI_ISL_455670\|2020-05-0 | ···················· | ···················· | ···················· | ···················· | ··A················· | ···················· |
| C | 3746 | hCoV-19/India/WB-S51/2020\|EPI_ISL_455671\|2020-05-0 | ···················· | ···················· | ···················· | ···················· | ···················· | ···················· |
| C | 3747 | hCoV-19/India/WB-S52/2020\|EPI_ISL_455672\|2020-05-0 | ···················· | ···················· | ···················· | ···················· | ···················· | ···················· |
| C | 3748 | hCoV-19/India/WB-S53/2020\|EPI_ISL_455673\|2020-05-0 | ···················· | ···················· | ···················· | ···················· | ···················· | ···················· |
| C | 3749 | hCoV-19/India/WB-S55/2020\|EPI_ISL_455674\|2020-05-0 | ···················· | ···················· | ···················· | ···················· | ···················· | ···················· |
| C | 3750 | hCoV-19/India/WB-S56/2020\|EPI_ISL_455675\|2020-05-0 | ···················· | ···················· | ···················· | ···················· | ···················· | ···················· |
| C | 3751 | hCoV-19/India/WB-S58/2020\|EPI_ISL_455676\|2020-05-0 | ···················· | ···················· | ···················· | ···················· | ···················· | ···················· |
| C | 3752 | hCoV-19/India/WB-S61/2020\|EPI_ISL_455678\|2020-04-3 | ···················· | ···················· | ···················· | ···················· | ···················· | ···················· |

| | | | | | | | | | |
|---|---|---|---|---|---|---|---|---|---|
| C | 3753 | hCoV-19/India/WB-S62/2020\|EPI_ISL_455679\|2020-05-0 | .................... | .................... | .................... | .................... | .................... | .................... | |
| C | 3754 | hCoV-19/Changzhou/JS27/2020\|EPI_ISL_455680\|2020-01 | .................... | .................... | .................... | .................... | .................... | .................... | |
| C | 3755 | hCoV-19/env/USA/UN-UF-12/2020\|EPI_ISL_455682\|2020- | .................... | .................... | .................... | .................... | .................... | .................... | |
| C | 3756 | hCoV-19/Zhejiang/CN1/2020\|EPI_ISL_455683\|2020-01-2 | .................... | .................... | .................... | .................... | .................... | .................... | |
| C | 3757 | hCoV-19/Zhejiang/CN2/2020\|EPI_ISL_455684\|2020-01-2 | .................... | .................... | .................... | .................... | .................... | .................... | |
| C | 3758 | hCoV-19/Zhejiang/CN5/2020\|EPI_ISL_455685\|2020-01-2 | .................... | .................... | .................... | .................... | .................... | .................... | |
| C | 3759 | hCoV-19/Zhejiang/CN3/2020\|EPI_ISL_455686\|2020-01-2 | .................... | .................... | .................... | .................... | .................... | .................... | |
| C | 3760 | hCoV-19/Zhejiang/CN4/2020\|EPI_ISL_455687\|2020-01-2 | .................... | .................... | .................... | .................... | .................... | .................... | |
| C | 3761 | hCoV-19/Zhejiang/OS1/2020\|EPI_ISL_455688\|2020-03-2 | .................... | .................... | .................... | .................... | .................... | .................... | |
| C | 3762 | hCoV-19/Zhejiang/OS5/2020\|EPI_ISL_455689\|2020-03-2 | .................... | .................... | .................... | .................... | .................... | .................... | |
| C | 3763 | hCoV-19/Zhejiang/OS6/2020\|EPI_ISL_455690\|2020-03-2 | .................... | .................... | .................... | .................... | .................... | .................... | |
| C | 3764 | hCoV-19/Zhejiang/OS3/2020\|EPI_ISL_455691\|2020-03-2 | .................... | .................... | .................... | .................... | .................... | .................... | |
| C | 3765 | hCoV-19/Zhejiang/OS2/2020\|EPI_ISL_455692\|2020-03-2 | .................... | .................... | .................... | .................... | .................... | .................... | |
| C | 3766 | hCoV-19/Zhejiang/OS4/2020\|EPI_ISL_455693\|2020-03-2 | .................... | .................... | .................... | .................... | .................... | .................... | |
| C | 3767 | hCoV-19/Vietnam/VNHN_1098/2020\|EPI_ISL_455694\|2020 | .................... | .................... | .................... | .................... | .................... | .................... | |
| C | 3768 | hCoV-19/Vietnam/VNHN_1099/2020\|EPI_ISL_455695\|2020 | .................... | .................... | .................... | .................... | .................... | .................... | |
| C | 3769 | hCoV-19/Vietnam/VNHN_1167/2020\|EPI_ISL_455696\|2020 | .................... | .................... | .................... | .................... | .................... | .................... | |
| C | 3770 | hCoV-19/Vietnam/VNHN_1528/2020\|EPI_ISL_455697\|2020 | .................... | .................... | .................... | .................... | .................... | .................... | |
| C | 3771 | hCoV-19/Vietnam/VNHN_1226/2020\|EPI_ISL_455698\|2020 | .................... | .................... | .................... | .................... | .................... | .................... | |
| C | 3772 | hCoV-19/Vietnam/VNHN_1492/2020\|EPI_ISL_455699\|2020 | .................... | .................... | .................... | .................... | .................... | .................... | |
| C | 3773 | hCoV-19/Scotland/EDB285/2020\|EPI_ISL_456901\|2020-0 | .................... | .................... | .................... | .................... | .................... | .................... | |
| C | 3774 | hCoV-19/Scotland/EDB289/2020\|EPI_ISL_456902\|2020-0 | .................... | .................... | .................... | .................... | .................... | .................... | |
| C | 3775 | hCoV-19/Scotland/EDB290/2020\|EPI_ISL_456903\|2020-0 | .................... | .................... | .................... | .................... | .................... | .................... | |
| C | 3776 | hCoV-19/Scotland/EDB2900/2020\|EPI_ISL_456904\|2020- | .................... | ...................A | .................... | .................... | .................... | .................... | 1 |
| C | 3777 | hCoV-19/Scotland/EDB291/2020\|EPI_ISL_456905\|2020-0 | .................... | .................... | .................... | .................... | .................... | .................... | |
| C | 3778 | hCoV-19/Scotland/EDB295/2020\|EPI_ISL_456906\|2020-0 | .................... | .................... | .................... | .................... | .................... | .................... | |
| C | 3779 | hCoV-19/Scotland/EDB2966/2020\|EPI_ISL_456908\|2020- | .................... | .................... | .................... | .................... | .................... | .................... | |
| C | 3781 | hCoV-19/Scotland/EDB5041/2020\|EPI_ISL_456953\|2020- | .................... | .................... | .................... | .................... | .................... | .................... | |
| C | 3782 | hCoV-19/Scotland/EDB5046/2020\|EPI_ISL_456954\|2020- | .................... | ...................A | .................... | .................... | .................... | .................... | 1 |
| C | 3783 | hCoV-19/Scotland/EDB5051/2020\|EPI_ISL_456955\|2020- | .................... | .................... | .................... | .................... | .................... | .................... | |
| C | 3784 | hCoV-19/Scotland/EDB5052/2020\|EPI_ISL_456956\|2020- | .................... | .................... | .................... | .................... | .................... | .................... | |
| C | 3785 | hCoV-19/Scotland/EDB5053/2020\|EPI_ISL_456957\|2020- | .................... | .................... | .................... | .................... | .................... | .................... | |
| C | 3786 | hCoV-19/Scotland/EDB5056/2020\|EPI_ISL_456958\|2020- | .................... | .................... | .................... | .................... | .................... | .................... | |
| C | 3787 | hCoV-19/Scotland/EDB5060/2020\|EPI_ISL_456960\|2020- | .................... | .................... | .................... | .................... | .................... | .................... | |
| C | 3788 | hCoV-19/Scotland/EDB5066/2020\|EPI_ISL_456961\|2020- | .................... | .................... | .................... | .................... | .................... | .................... | |
| C | 3789 | hCoV-19/Scotland/EDB5070/2020\|EPI_ISL_456962\|2020- | .................... | .................... | .................... | .................... | .................... | .................... | |
| C | 3790 | hCoV-19/Scotland/EDB5088/2020\|EPI_ISL_456964\|2020- | .................... | .................... | .................... | .................... | .................... | .................... | |
| C | 3791 | hCoV-19/Scotland/EDB5090/2020\|EPI_ISL_456965\|2020- | .................... | .................... | .................... | .................... | .................... | .................... | |
| C | 3792 | hCoV-19/Scotland/EDB5091/2020\|EPI_ISL_456966\|2020- | .................... | .................... | .................... | .................... | .................... | .................... | |
| C | 3793 | hCoV-19/Scotland/EDB5094/2020\|EPI_ISL_456967\|2020- | .................... | .................... | .................... | .................... | .................... | .................... | |
| C | 3794 | hCoV-19/Scotland/EDB5103/2020\|EPI_ISL_456973\|2020- | .................... | .................... | .................... | .................... | .................... | .................... | |
| C | 3795 | hCoV-19/Scotland/EDB5123/2020\|EPI_ISL_456974\|2020- | .................... | .A.................. | .................... | .................... | .................... | .................... | |
| C | 3796 | hCoV-19/Scotland/EDB5125/2020\|EPI_ISL_456975\|2020- | .................... | .................... | .................... | .................... | .................... | .................... | |
| C | 3797 | hCoV-19/Scotland/EDB5126/2020\|EPI_ISL_456976\|2020- | .................... | .A.................. | .................... | .................... | .................... | .................... | |
| C | 3798 | hCoV-19/Scotland/EDB5128/2020\|EPI_ISL_456977\|2020- | .................... | .A.................. | .................... | .................... | .................... | .................... | |
| C | 3799 | hCoV-19/Scotland/EDB5151/2020\|EPI_ISL_456978\|2020- | .................... | .A.................. | .................... | .................... | .................... | .................... | |
| C | 3800 | hCoV-19/Scotland/EDB5155/2020\|EPI_ISL_456979\|2020- | .................... | .A.................. | .................... | .................... | .................... | .................... | |
| C | 3801 | hCoV-19/Scotland/EDB5181/2020\|EPI_ISL_456984\|2020- | .................... | .................... | .................... | .................... | .................... | .................... | |
| C | 3802 | hCoV-19/Scotland/EDB5184/2020\|EPI_ISL_456985\|2020- | .................... | .................... | .................... | .................... | .................... | .................... | |
| C | 3803 | hCoV-19/Scotland/EDB5209/2020\|EPI_ISL_456986\|2020- | .................... | .A.................. | .................... | .................... | .................... | .................... | |
| C | 3804 | hCoV-19/Scotland/EDB5214/2020\|EPI_ISL_456987\|2020- | .................... | .A.................. | .................... | .................... | .................... | .................... | |
| C | 3805 | hCoV-19/Scotland/EDB5218/2020\|EPI_ISL_456988\|2020- | .................... | .................... | .................... | .................... | .................... | .................... | |
| C | 3806 | hCoV-19/Scotland/EDB5220/2020\|EPI_ISL_456989\|2020- | .................... | .................... | .................... | .................... | .................... | .................... | |
| C | 3808 | hCoV-19/Scotland/EDB5245/2020\|EPI_ISL_456992\|2020- | .................... | .................... | .................... | .................... | .................... | .................... | |
| C | 3810 | hCoV-19/Scotland/EDB5249/2020\|EPI_ISL_456994\|2020- | .................... | .A.................. | .................... | .................... | .................... | .................... | |
| C | 3812 | hCoV-19/Scotland/EDB5251/2020\|EPI_ISL_456996\|2020- | .................... | .................... | .................... | .................... | .................... | .................... | |
| C | 3813 | hCoV-19/Scotland/EDB5276/2020\|EPI_ISL_456997\|2020- | .................... | .................... | .................... | .................... | .................... | .................... | |
| C | 3814 | hCoV-19/Scotland/EDB5280/2020\|EPI_ISL_456998\|2020- | .................... | .................... | .................... | .................... | .................... | .................... | |
| C | 3816 | hCoV-19/Northern Ireland/NIRE-FC8F0/2020\|EPI_ISL_4 | .................... | .................... | .................... | .................... | .................... | .................... | |
| C | 3817 | hCoV-19/Northern Ireland/NIRE-FBFFC/2020\|EPI_ISL_4 | .................... | .................... | .................... | .................... | .................... | .................... | |
| C | 3818 | hCoV-19/Northern Ireland/NIRE-FBF83/2020\|EPI_ISL_4 | .................... | .................... | .................... | .................... | .................... | .................... | |
| C | 3819 | hCoV-19/Northern Ireland/NIRE-FC6BA/2020\|EPI_ISL_4 | .................... | .................... | .................... | .................... | .................... | .................... | |
| C | 3820 | hCoV-19/USA/MA-MGH-00285/2020\|EPI_ISL_460169\|2020- | .................... | .................... | .................... | .................... | .................... | .................... | |
| C | 3821 | hCoV-19/USA/MA-MGH-00145/2020\|EPI_ISL_460170\|2020- | .................... | .................... | .................... | .................... | .................... | .................... | |
| C | 3822 | hCoV-19/USA/MA-MGH-00291/2020\|EPI_ISL_460171\|2020- | .................... | .................... | .................... | .................... | .................... | .................... | |
| C | 3823 | hCoV-19/USA/MA-MGH-00276/2020\|EPI_ISL_460172\|2020- | .................... | .................... | .................... | .................... | .................... | .................... | |
| C | 3824 | hCoV-19/USA/MA-MGH-00254/2020\|EPI_ISL_460173\|2020- | .................... | .................... | .................... | .................... | .................... | .................... | |
| C | 3825 | hCoV-19/USA/MA-MGH-00059/2020\|EPI_ISL_460174\|2020- | .................... | .................... | .................... | .................... | .................... | .................... | |
| C | 3826 | hCoV-19/USA/MA-MGH-00055/2020\|EPI_ISL_460175\|2020- | .................... | .................... | .................... | .................... | .................... | .................... | |
| C | 3827 | hCoV-19/Scotland/CVR837/2020\|EPI_ISL_461705\|2020-0 | .................... | .................... | .................... | .................... | .................... | .................... | |
| C | 3828 | hCoV-19/Scotland/EDB286/2020\|EPI_ISL_461706\|2020-0 | .................... | .................... | .................... | .................... | .................... | .................... | |
| C | 3829 | hCoV-19/Scotland/EDB294/2020\|EPI_ISL_461707\|2020-0 | .................... | .................... | .................... | .................... | .................... | .................... | |
| C | 3830 | hCoV-19/Scotland/EDB306/2020\|EPI_ISL_461708\|2020-0 | .................... | ...................A | .................... | .................... | .................... | .................... | 1 |
| C | 3831 | hCoV-19/Scotland/EDB375/2020\|EPI_ISL_461710\|2020-0 | .................... | .................... | .................... | .................... | .................... | .................... | |
| C | 3832 | hCoV-19/Scotland/EDB4454/2020\|EPI_ISL_461711\|2020- | .................... | .................... | .................... | .................... | .................... | .................... | |
| C | 3833 | hCoV-19/Scotland/EDB4455/2020\|EPI_ISL_461712\|2020- | .................... | .A.................. | .................... | .................... | .................... | .................... | |
| C | 3834 | hCoV-19/Scotland/EDB4457/2020\|EPI_ISL_461713\|2020- | .................... | .................... | .................... | .................... | .................... | .................... | |
| C | 3835 | hCoV-19/Scotland/EDB4517/2020\|EPI_ISL_461714\|2020- | .................... | .................... | .................... | .................... | .................... | .................... | |

| | | | | | | | |
|---|---|---|---|---|---|---|---|
| C | 3836 hCoV-19/Scotland/EDB4533/2020\|EPI_ISL_461716\|2020- | ···················· | ·A·················· | ···················· | ···················· | ···················· | ···················· |
| C | 3837 hCoV-19/Scotland/EDB4535/2020\|EPI_ISL_461718\|2020- | ···················· | ···················· | ···················· | ···················· | ···················· | ···················· |
| C | 3838 hCoV-19/Scotland/EDB4536/2020\|EPI_ISL_461719\|2020- | ···················· | ···················· | ···················· | ···················· | ···················· | ···················· |
| C | 3839 hCoV-19/Scotland/EDB4557/2020\|EPI_ISL_461725\|2020- | ···················· | ···················· | ···················· | ···················· | ···················· | ···················· |
| C | 3840 hCoV-19/Scotland/EDB4559/2020\|EPI_ISL_461727\|2020- | ···················· | ···················· | ···················· | ···················· | ···················· | ···················· |
| C | 3841 hCoV-19/England/NOTT-110DBA/2020\|EPI_ISL_448417\|20 | ···················· | ···················· | ···················· | ···················· | ···················· | ···················· |
| C | 3842 hCoV-19/England/NOTT-110DC9/2020\|EPI_ISL_448418\|20 | ···················· | ···················· | ···················· | ···················· | ···················· | ···················· |
| C | 3843 hCoV-19/England/NOTT-110DE7/2020\|EPI_ISL_448420\|20 | ···················· | ···················· | ···················· | ···················· | ···················· | ···················· |
| C | 3844 hCoV-19/England/NOTT-110DF6/2020\|EPI_ISL_448421\|20 | ···················· | ···················· | ···················· | ···················· | ···················· | ···················· |
| C | 3845 hCoV-19/England/NOTT-110E99/2020\|EPI_ISL_448430\|20 | ···················· | ···················· | ···················· | ···················· | ···················· | ···················· |
| C | 3847 hCoV-19/England/NOTT-110F1E/2020\|EPI_ISL_448436\|20 | ···················· | ···················· | ···················· | ···················· | ···················· | ···················· |
| C | 3848 hCoV-19/England/NOTT-110F3C/2020\|EPI_ISL_448438\|20 | ···················· | ···················· | ···················· | ···················· | ···················· | ···················· |
| C | 3849 hCoV-19/England/NOTT-110F69/2020\|EPI_ISL_448440\|20 | ···················· | ···················· | ···················· | ···················· | ···················· | ···················· |
| C | 3850 hCoV-19/England/NOTT-110F78/2020\|EPI_ISL_448441\|20 | ···················· | ···················· | ···················· | ···················· | ···················· | ···················· |
| C | 3851 hCoV-19/England/NOTT-110F96/2020\|EPI_ISL_448443\|20 | ···················· | ···················· | ···················· | ···················· | ···················· | ···················· |
| C | 3852 hCoV-19/England/NOTT-110FA5/2020\|EPI_ISL_448444\|20 | ···················· | ···················· | ···················· | ···················· | ···················· | ···················· |
| C | 3853 hCoV-19/England/NOTT-110FB4/2020\|EPI_ISL_448445\|20 | ···················· | ···················· | ···················· | ···················· | ···················· | ···················· |
| C | 3854 hCoV-19/England/NOTT-110FC3/2020\|EPI_ISL_448446\|20 | ···················· | ···················· | ···················· | ···················· | ···················· | ···················· |
| C | 3855 hCoV-19/England/NOTT-110FE1/2020\|EPI_ISL_448448\|20 | ···················· | ···················· | ···················· | ···················· | ···················· | ···················· |
| C | 3856 hCoV-19/England/OXON-AC428/2020\|EPI_ISL_448449\|202 | ···················· | ···················· | ···················· | ···················· | ···················· | ···················· |
| C | 3857 hCoV-19/England/OXON-AC437/2020\|EPI_ISL_448450\|202 | ···················· | ···················· | ···················· | ···················· | ···················· | ···················· |
| C | 3858 hCoV-19/England/OXON-AC446/2020\|EPI_ISL_448451\|202 | ···················· | ···················· | ···················· | ···················· | ···················· | ···················· |
| C | 3859 hCoV-19/England/OXON-AC491/2020\|EPI_ISL_448453\|202 | ···················· | ·············A······ | ···················· | ···················· | ···················· | ···················· |
| C | 3860 hCoV-19/England/OXON-AC4A0/2020\|EPI_ISL_448454\|202 | ···················· | ···················· | ···················· | ···················· | ···················· | ···················· |
| C | 3861 hCoV-19/England/OXON-AC4BF/2020\|EPI_ISL_448455\|202 | ···················· | ···················· | ···················· | ···················· | ···················· | ···················· |
| C | 3862 hCoV-19/England/OXON-AC4CE/2020\|EPI_ISL_448456\|202 | ···················· | ···················· | ···················· | ···················· | ···················· | ···················· |
| C | 3863 hCoV-19/England/OXON-AC507/2020\|EPI_ISL_448457\|202 | ···················· | ···················· | ···················· | ···················· | ···················· | ···················· |
| C | 3864 hCoV-19/England/OXON-AC534/2020\|EPI_ISL_448458\|202 | ···················· | ···················· | ···················· | ···················· | ···················· | ···················· |
| C | 3865 hCoV-19/England/OXON-AC543/2020\|EPI_ISL_448459\|202 | ···················· | ···················· | ···················· | ···················· | ···················· | ···················· |
| C | 3866 hCoV-19/England/OXON-AC798/2020\|EPI_ISL_448483\|202 | ···················· | ···················· | ···················· | ···················· | ···················· | ···················· |
| C | 3867 hCoV-19/England/OXON-AC81D/2020\|EPI_ISL_448486\|202 | ···················· | ···················· | ···················· | ···················· | ···················· | ···················· |
| C | 3868 hCoV-19/England/OXON-AC83B/2020\|EPI_ISL_448487\|202 | ···················· | ···················· | ···················· | ···················· | ···················· | ···················· |
| C | 3869 hCoV-19/England/OXON-AC868/2020\|EPI_ISL_448488\|202 | ···················· | ···················· | ···················· | ···················· | ···················· | ···················· |
| C | 3870 hCoV-19/Belgium/ITM_C184/2020\|EPI_ISL_450730\|2020- | ···················· | ···················· | ···················· | ···················· | ···················· | ···················· |
| C | 3871 hCoV-19/Belgium/ITM_C193/2020\|EPI_ISL_450731\|2020- | ···················· | ···················· | ···················· | ···················· | ···················· | ···················· |
| C | 3872 hCoV-19/Belgium/ITM_C237/2020\|EPI_ISL_450732\|2020- | ···················· | ···················· | ···················· | ···················· | ···················· | ···················· |
| C | 3873 hCoV-19/Belgium/ITM_C338/2020\|EPI_ISL_450734\|2020- | ···················· | ···················· | ···················· | ···················· | ···················· | ···················· |
| C | 3874 hCoV-19/Belgium/ITM_C376/2020\|EPI_ISL_450736\|2020- | ···················· | ···················· | ···················· | ···················· | ···················· | ···················· |
| C | 3875 hCoV-19/Belgium/ITM_C413/2020\|EPI_ISL_450737\|2020- | ···················· | ···················· | ···················· | ···················· | ···················· | ···················· |
| C | 3876 hCoV-19/Vietnam/HCMC-05004/2020\|EPI_ISL_450738\|202 | ···················· | ···················· | ···················· | ···················· | ···················· | ···················· |
| C | 3877 hCoV-19/Vietnam/HCMC-35005/2020\|EPI_ISL_450739\|202 | ···················· | ···················· | ···················· | ···················· | ···················· | ···················· |
| C | 3878 hCoV-19/Vietnam/HCMC-35011/2020\|EPI_ISL_450740\|202 | ···················· | ···················· | ···················· | ···················· | ···················· | ···················· |
| C | 3879 hCoV-19/Vietnam/HCMC-35016/2020\|EPI_ISL_450741\|202 | ···················· | ···················· | ···················· | ···················· | ···················· | ···················· |
| C | 3880 hCoV-19/Vietnam/HCMC-35018/2020\|EPI_ISL_450742\|202 | ···················· | ···················· | ···················· | ···················· | ···················· | ···················· |
| C | 3881 hCoV-19/Vietnam/HCMC-35019/2020\|EPI_ISL_450743\|202 | ···················· | ···················· | ···················· | ···················· | ···················· | ···················· |
| C | 3882 hCoV-19/Vietnam/HCMC-35020/2020\|EPI_ISL_450744\|202 | ···················· | ···················· | ···················· | ···················· | ···················· | ···················· |
| C | 3883 hCoV-19/Vietnam/HCMC-35021/2020\|EPI_ISL_450745\|202 | ···················· | ···················· | ···················· | ···················· | ···················· | ···················· |
| C | 3884 hCoV-19/Poland/Pom2/2020\|EPI_ISL_450746\|2020-05-08 | ···················· | ···················· | ···················· | ···················· | ······T············· | ···················· |
| C | 3885 hCoV-19/Canada/ON_SHSC-01/2020\|EPI_ISL_450747\|2020 | ···················· | ···················· | ···················· | ···················· | ···················· | ···················· |
| C | 3886 hCoV-19/USA/MN-MDH-274/2020\|EPI_ISL_450748\|2020-03 | ···················· | ···················· | ···················· | ···················· | ···················· | ···················· |
| C | 3887 hCoV-19/USA/MN-MDH-275/2020\|EPI_ISL_450749\|2020-03 | ···················· | ···················· | ···················· | ···················· | ···················· | ···················· |
| C | 3888 hCoV-19/USA/MN-MDH-276/2020\|EPI_ISL_450750\|2020-03 | ···················· | ···················· | ···················· | ···················· | ···················· | ···················· |
| C | 3889 hCoV-19/USA/MN-MDH-277/2020\|EPI_ISL_450751\|2020-03 | ···················· | ···················· | ···················· | ···················· | ···················· | ···················· |
| C | 3890 hCoV-19/USA/MN-MDH-278/2020\|EPI_ISL_450752\|2020-03 | ···················· | ···················· | ···················· | ···················· | ···················· | ···················· |
| C | 3891 hCoV-19/USA/MN-MDH-282/2020\|EPI_ISL_450753\|2020-03 | ···················· | ···················· | ···················· | ···················· | ···················· | ···················· |
| C | 3892 hCoV-19/USA/MN-MDH-283/2020\|EPI_ISL_450754\|2020-03 | ···················· | ···················· | ···················· | ···················· | ···················· | ···················· |
| C | 3893 hCoV-19/USA/MN-MDH-285/2020\|EPI_ISL_450755\|2020-03 | ···················· | ···················· | ···················· | ···················· | ···················· | ···················· |
| C | 3894 hCoV-19/USA/MN-MDH-287/2020\|EPI_ISL_450756\|2020-03 | ···················· | ····G··············· | ···················· | ···················· | ···················· | ···················· |
| C | 3895 hCoV-19/USA/MN-MDH-288/2020\|EPI_ISL_450757\|2020-03 | ···················· | ····G··············· | ···················· | ···················· | ···················· | ···················· |
| C | 3896 hCoV-19/USA/MN-MDH-289/2020\|EPI_ISL_450758\|2020-03 | ···················· | ····G··············· | ···················· | ···················· | ···················· | ···················· |
| C | 3897 hCoV-19/USA/MN-MDH-290/2020\|EPI_ISL_450759\|2020-03 | ···················· | ··········T········· | ···················· | ···················· | ···················· | ···················· |
| C | 3898 hCoV-19/USA/MN-MDH-291/2020\|EPI_ISL_450760\|2020-03 | ···················· | ···················· | ···················· | ···················· | ···················· | ···················· |
| C | 3899 hCoV-19/USA/MN-MDH-292/2020\|EPI_ISL_450761\|2020-03 | ···················· | ···················· | ···················· | ···················· | ···················· | ···················· |
| C | 3900 hCoV-19/USA/MN-MDH-293/2020\|EPI_ISL_450762\|2020-03 | ···················· | ···················· | ···················· | ···················· | ···················· | ···················· |
| C | 3901 hCoV-19/USA/MN-MDH-295/2020\|EPI_ISL_450763\|2020-03 | ···················· | ···················· | ···················· | ···················· | ···················· | ···················· |
| C | 3902 hCoV-19/USA/MN-MDH-296/2020\|EPI_ISL_450764\|2020-03 | ···················· | ···················· | ···················· | ···················· | ···················· | ···················· |
| C | 3903 hCoV-19/USA/MN-MDH-297/2020\|EPI_ISL_450765\|2020-03 | ···················· | ···················· | ···················· | ···················· | ···················· | ···················· |
| C | 3904 hCoV-19/USA/MN-MDH-299/2020\|EPI_ISL_450766\|2020-03 | ···················· | ···················· | ···················· | ···················· | ···················· | ···················· |
| C | 3905 hCoV-19/USA/MN-MDH-300/2020\|EPI_ISL_450767\|2020-03 | ···················· | ···················· | ···················· | ···················· | ···················· | ···················· |
| C | 3906 hCoV-19/USA/MN-MDH-301/2020\|EPI_ISL_450768\|2020-03 | ···················· | ···················· | ···················· | ···················· | ···················· | ···················· |
| C | 3907 hCoV-19/USA/MN-MDH-304/2020\|EPI_ISL_450769\|2020-03 | ···················· | ····G··············· | ···················· | ···················· | ···················· | ···················· |
| C | 3908 hCoV-19/USA/MN-MDH-305/2020\|EPI_ISL_450770\|2020-03 | ···················· | ···················· | ···················· | ···················· | ···················· | ···················· |
| C | 3909 hCoV-19/Australia/VPRL010/2020\|EPI_ISL_455602\|2020 | ···················· | ···················· | ···················· | ···················· | ···················· | ···················· |
| C | 3910 hCoV-19/Australia/VPRL014/2020\|EPI_ISL_455603\|2020 | ···················· | ···················· | ···················· | ···················· | ···················· | ···················· |
| C | 3911 hCoV-19/Thailand/Bangkok_2420/2020\|EPI_ISL_455604\| | ···················· | ···················· | ···················· | ···················· | ···················· | ···················· |
| C | 3912 hCoV-19/Thailand/Nonthaburi_2472/2020\|EPI_ISL_4556 | ···················· | ···················· | ···················· | ···················· | ···················· | ···················· |
| C | 3913 hCoV-19/Thailand/Bangkok_2488/2020\|EPI_ISL_455606\| | ···················· | ···················· | ···················· | ···················· | ···················· | ···················· |
| C | 3914 hCoV-19/Thailand/Bangkok_2489/2020\|EPI_ISL_455607\| | ···················· | ···················· | ···················· | ···················· | ···················· | ···················· |

<table>
<tr><td>C</td><td>3915</td><td>hCoV-19/Thailand/Bangkok_2504/2020|EPI_ISL_455608|</td><td>....................</td><td>....................</td><td>.....................</td><td>....................</td><td>....................</td><td>....................</td><td></td></tr>
<tr><td>C</td><td>3916</td><td>hCoV-19/USA/LA-BIE-051/2020|EPI_ISL_455609|2020-04</td><td>....................</td><td>....................</td><td>.....................</td><td>....................</td><td>....................</td><td>....................</td><td></td></tr>
<tr><td>C</td><td>3917</td><td>hCoV-19/USA/LA-BIE-054/2020|EPI_ISL_455610|2020-04</td><td>....................</td><td>....................</td><td>.....................</td><td>....................</td><td>....................</td><td>....................</td><td></td></tr>
<tr><td>C</td><td>3918</td><td>hCoV-19/USA/LA-BIE-055/2020|EPI_ISL_455611|2020-04</td><td>....................</td><td>....................</td><td>.....................</td><td>....................</td><td>....................</td><td>....................</td><td></td></tr>
<tr><td>C</td><td>3919</td><td>hCoV-19/USA/LA-BIE-056/2020|EPI_ISL_455612|2020-04</td><td>....................</td><td>....................</td><td>.....................</td><td>....................</td><td>....................</td><td>....................</td><td></td></tr>
<tr><td>C</td><td>3920</td><td>hCoV-19/USA/LA-BIE-058/2020|EPI_ISL_455613|2020-04</td><td>....................</td><td>....................</td><td>.....................</td><td>....................</td><td>....................</td><td>....................</td><td></td></tr>
<tr><td>C</td><td>3921</td><td>hCoV-19/USA/LA-BIE-059/2020|EPI_ISL_455614|2020-04</td><td>....................</td><td>....................</td><td>.....................</td><td>....................</td><td>....................</td><td>....................</td><td></td></tr>
<tr><td>C</td><td>3922</td><td>hCoV-19/USA/LA-BIE-061/2020|EPI_ISL_455615|2020-04</td><td>....................</td><td>....................</td><td>.....................</td><td>....................</td><td>....................</td><td>....................</td><td></td></tr>
<tr><td>C</td><td>3923</td><td>hCoV-19/USA/LA-BIE-062/2020|EPI_ISL_455616|2020-04</td><td>....................</td><td>....................</td><td>.....................</td><td>....................</td><td>....................</td><td>....................</td><td></td></tr>
<tr><td>C</td><td>3924</td><td>hCoV-19/USA/LA-BIE-063/2020|EPI_ISL_455617|2020-04</td><td>....................</td><td>....................</td><td>.....................</td><td>....................</td><td>....................</td><td>....................</td><td></td></tr>
<tr><td>C</td><td>3925</td><td>hCoV-19/USA/LA-BIE-064/2020|EPI_ISL_455618|2020-04</td><td>....................</td><td>....................</td><td>.....................</td><td>....................</td><td>....................</td><td>....................</td><td></td></tr>
<tr><td>C</td><td>3926</td><td>hCoV-19/USA/LA-BIE-065/2020|EPI_ISL_455619|2020-04</td><td>....................</td><td>....................</td><td>.....................</td><td>....................</td><td>....................</td><td>....................</td><td></td></tr>
<tr><td>C</td><td>3927</td><td>hCoV-19/USA/LA-BIE-066/2020|EPI_ISL_455620|2020-04</td><td>....................</td><td>....................</td><td>.....................</td><td>....................</td><td>....................</td><td>....................</td><td></td></tr>
<tr><td>C</td><td>3928</td><td>hCoV-19/USA/LA-BIE-068/2020|EPI_ISL_455621|2020-04</td><td>....................</td><td>....................</td><td>.....................</td><td>....................</td><td>....................</td><td>....................</td><td></td></tr>
<tr><td>C</td><td>3929</td><td>hCoV-19/Scotland/EDB3058/2020|EPI_ISL_456910|2020-</td><td>....................</td><td>....................</td><td>.....................</td><td>....................</td><td>....................</td><td>....................</td><td></td></tr>
<tr><td>C</td><td>3930</td><td>hCoV-19/Scotland/EDB3062/2020|EPI_ISL_456911|2020-</td><td>....................</td><td>....................</td><td>.....................</td><td>....................</td><td>....................</td><td>....................</td><td></td></tr>
<tr><td>C</td><td>3931</td><td>hCoV-19/Scotland/EDB3105/2020|EPI_ISL_456913|2020-</td><td>....................</td><td>....................</td><td>.....................</td><td>....................</td><td>....................</td><td>....................</td><td></td></tr>
<tr><td>C</td><td>3932</td><td>hCoV-19/Scotland/EDB3295/2020|EPI_ISL_456914|2020-</td><td>....................</td><td>...................A</td><td>.....................</td><td>....................</td><td>....................</td><td>....................</td><td>1</td></tr>
<tr><td>C</td><td>3934</td><td>hCoV-19/Scotland/EDB4009/2020|EPI_ISL_456916|2020-</td><td>....................</td><td>....................</td><td>.....................</td><td>....................</td><td>....................</td><td>....................</td><td></td></tr>
<tr><td>C</td><td>3936</td><td>hCoV-19/Scotland/EDB4146/2020|EPI_ISL_456920|2020-</td><td>....................</td><td>...................A</td><td>.....................</td><td>....................</td><td>....................</td><td>....................</td><td>1</td></tr>
<tr><td>C</td><td>3937</td><td>hCoV-19/Scotland/EDB4149/2020|EPI_ISL_456921|2020-</td><td>....................</td><td>.A..................</td><td>.....................</td><td>....................</td><td>....................</td><td>....................</td><td></td></tr>
<tr><td>C</td><td>3938</td><td>hCoV-19/Scotland/EDB4150/2020|EPI_ISL_456922|2020-</td><td>....................</td><td>.A..................</td><td>.....................</td><td>....................</td><td>....................</td><td>....................</td><td></td></tr>
<tr><td>C</td><td>3939</td><td>hCoV-19/Scotland/EDB4152/2020|EPI_ISL_456923|2020-</td><td>....................</td><td>.A..................</td><td>.....................</td><td>....................</td><td>....................</td><td>....................</td><td></td></tr>
<tr><td>C</td><td>3940</td><td>hCoV-19/Scotland/EDB4153/2020|EPI_ISL_456924|2020-</td><td>....................</td><td>...................A</td><td>.....................</td><td>....................</td><td>....................</td><td>....................</td><td>1</td></tr>
<tr><td>C</td><td>3941</td><td>hCoV-19/Scotland/EDB4157/2020|EPI_ISL_456925|2020-</td><td>....................</td><td>....................</td><td>.....................</td><td>....................</td><td>....................</td><td>....................</td><td></td></tr>
<tr><td>C</td><td>3942</td><td>hCoV-19/Scotland/EDB4158/2020|EPI_ISL_456926|2020-</td><td>....................</td><td>...................A</td><td>.....................</td><td>....................</td><td>....................</td><td>....................</td><td>1</td></tr>
<tr><td>C</td><td>3943</td><td>hCoV-19/Scotland/EDB4160/2020|EPI_ISL_456927|2020-</td><td>....................</td><td>...................A</td><td>.....................</td><td>....................</td><td>....................</td><td>....................</td><td>1</td></tr>
<tr><td>C</td><td>3944</td><td>hCoV-19/Scotland/EDB4161/2020|EPI_ISL_456928|2020-</td><td>....................</td><td>....................</td><td>.....................</td><td>....................</td><td>....................</td><td>....................</td><td></td></tr>
<tr><td>C</td><td>3945</td><td>hCoV-19/Scotland/EDB4168/2020|EPI_ISL_456930|2020-</td><td>....................</td><td>...................A</td><td>.....................</td><td>....................</td><td>....................</td><td>....................</td><td>1</td></tr>
<tr><td>C</td><td>3946</td><td>hCoV-19/Scotland/EDB4169/2020|EPI_ISL_456931|2020-</td><td>....................</td><td>....................</td><td>.....................</td><td>....................</td><td>....................</td><td>....................</td><td></td></tr>
<tr><td>C</td><td>3947</td><td>hCoV-19/Scotland/EDB4174/2020|EPI_ISL_456932|2020-</td><td>....................</td><td>....................</td><td>.....................</td><td>....................</td><td>....................</td><td>....................</td><td></td></tr>
<tr><td>C</td><td>3948</td><td>hCoV-19/Scotland/EDB4196/2020|EPI_ISL_456933|2020-</td><td>....................</td><td>....................</td><td>.....................</td><td>....................</td><td>....................</td><td>....................</td><td></td></tr>
<tr><td>C</td><td>3949</td><td>hCoV-19/Scotland/EDB4201/2020|EPI_ISL_456934|2020-</td><td>....................</td><td>....................</td><td>.....................</td><td>....................</td><td>....................</td><td>....................</td><td></td></tr>
<tr><td>C</td><td>3950</td><td>hCoV-19/Scotland/EDB4226/2020|EPI_ISL_456938|2020-</td><td>....................</td><td>.A..................</td><td>.....................</td><td>....................</td><td>....................</td><td>....................</td><td></td></tr>
<tr><td>C</td><td>3951</td><td>hCoV-19/Scotland/EDB4249/2020|EPI_ISL_456939|2020-</td><td>....................</td><td>....................</td><td>.....................</td><td>....................</td><td>....................</td><td>....................</td><td></td></tr>
<tr><td>C</td><td>3954</td><td>hCoV-19/Scotland/EDB4486/2020|EPI_ISL_456943|2020-</td><td>....................</td><td>....................</td><td>.....................</td><td>....................</td><td>....................</td><td>....................</td><td></td></tr>
<tr><td>C</td><td>3955</td><td>hCoV-19/Scotland/EDB4494/2020|EPI_ISL_456944|2020-</td><td>....................</td><td>....................</td><td>.....................</td><td>....................</td><td>....................</td><td>....................</td><td></td></tr>
<tr><td>C</td><td>3956</td><td>hCoV-19/Scotland/EDB4504/2020|EPI_ISL_456945|2020-</td><td>....................</td><td>.A..................</td><td>.....................</td><td>....................</td><td>....................</td><td>....................</td><td></td></tr>
<tr><td>C</td><td>3959</td><td>hCoV-19/Scotland/EDB4511/2020|EPI_ISL_456948|2020-</td><td>....................</td><td>....................</td><td>.....................</td><td>....................</td><td>....................</td><td>....................</td><td></td></tr>
<tr><td>C</td><td>3960</td><td>hCoV-19/Scotland/EDB4515/2020|EPI_ISL_456949|2020-</td><td>....................</td><td>.A..................</td><td>.....................</td><td>....................</td><td>....................</td><td>....................</td><td></td></tr>
<tr><td>C</td><td>3962</td><td>hCoV-19/Scotland/EDB5040/2020|EPI_ISL_456952|2020-</td><td>....................</td><td>....................</td><td>.....................</td><td>....................</td><td>....................</td><td>....................</td><td></td></tr>
<tr><td>C</td><td>3963</td><td>hCoV-19/Scotland/GCVR-172C8C/2020|EPI_ISL_459100|2</td><td>....................</td><td>....................</td><td>.....................</td><td>....................</td><td>....................</td><td>....................</td><td></td></tr>
<tr><td>C</td><td>3964</td><td>hCoV-19/Scotland/GCVR-173DC4/2020|EPI_ISL_459102|2</td><td>....................</td><td>....................</td><td>.....................</td><td>....................</td><td>....................</td><td>....................</td><td></td></tr>
<tr><td>C</td><td>3965</td><td>hCoV-19/Scotland/GCVR-173D2E/2020|EPI_ISL_459103|2</td><td>....................</td><td>....................</td><td>.....................</td><td>....................</td><td>....................</td><td>....................</td><td></td></tr>
<tr><td>C</td><td>3966</td><td>hCoV-19/Scotland/GCVR-173B8E/2020|EPI_ISL_459104|2</td><td>....................</td><td>....................</td><td>.....................</td><td>....................</td><td>....................</td><td>....................</td><td></td></tr>
<tr><td>C</td><td>3967</td><td>hCoV-19/Scotland/GCVR-173B15/2020|EPI_ISL_459105|2</td><td>....................</td><td>....................</td><td>.....................</td><td>....................</td><td>....................</td><td>....................</td><td></td></tr>
<tr><td>C</td><td>3968</td><td>hCoV-19/Scotland/GCVR-173CB8/2020|EPI_ISL_459106|2</td><td>....................</td><td>....................</td><td>.....................</td><td>....................</td><td>....................</td><td>....................</td><td></td></tr>
<tr><td>C</td><td>3969</td><td>hCoV-19/Scotland/GCVR-173B24/2020|EPI_ISL_459107|2</td><td>....................</td><td>....................</td><td>.....................</td><td>....................</td><td>....................</td><td>....................</td><td></td></tr>
<tr><td>C</td><td>3970</td><td>hCoV-19/Scotland/GCVR-172DD4/2020|EPI_ISL_459151|2</td><td>....................</td><td>....................</td><td>.....................</td><td>....................</td><td>....................</td><td>....................</td><td></td></tr>
<tr><td>C</td><td>3971</td><td>hCoV-19/Scotland/GCVR-173C9A/2020|EPI_ISL_459153|2</td><td>....................</td><td>....................</td><td>.....................</td><td>....................</td><td>....................</td><td>....................</td><td></td></tr>
<tr><td>C</td><td>3972</td><td>hCoV-19/Scotland/GCVR-172BAD/2020|EPI_ISL_459154|2</td><td>....................</td><td>....................</td><td>.....................</td><td>....................</td><td>....................</td><td>....................</td><td></td></tr>
<tr><td>C</td><td>3973</td><td>hCoV-19/Scotland/GCVR-173CA9/2020|EPI_ISL_459155|2</td><td>....................</td><td>....................</td><td>.....................</td><td>....................</td><td>....................</td><td>....................</td><td></td></tr>
<tr><td>C</td><td>3974</td><td>hCoV-19/Scotland/GCVR-172B8F/2020|EPI_ISL_459156|2</td><td>....................</td><td>....................</td><td>.....................</td><td>....................</td><td>....................</td><td>....................</td><td></td></tr>
<tr><td>C</td><td>3975</td><td>hCoV-19/Scotland/GCVR-172CAA/2020|EPI_ISL_459157|2</td><td>....................</td><td>....................</td><td>.....................</td><td>....................</td><td>....................</td><td>....................</td><td></td></tr>
<tr><td>C</td><td>3976</td><td>hCoV-19/Scotland/GCVR-173D00/2020|EPI_ISL_459158|2</td><td>....................</td><td>...........A........</td><td>.....................</td><td>....................</td><td>....................</td><td>....................</td><td></td></tr>
<tr><td>C</td><td>3977</td><td>hCoV-19/Scotland/GCVR-173B42/2020|EPI_ISL_459161|2</td><td>....................</td><td>....................</td><td>.....................</td><td>....................</td><td>....................</td><td>....................</td><td></td></tr>
<tr><td>C</td><td>3978</td><td>hCoV-19/Scotland/GCVR-172C5F/2020|EPI_ISL_459164|2</td><td>....................</td><td>....................</td><td>.....................</td><td>....................</td><td>....................</td><td>....................</td><td></td></tr>
<tr><td>C</td><td>3979</td><td>hCoV-19/Scotland/GCVR-173D1F/2020|EPI_ISL_459165|2</td><td>....................</td><td>...........A........</td><td>.....................</td><td>....................</td><td>....................</td><td>....................</td><td></td></tr>
<tr><td>C</td><td>3980</td><td>hCoV-19/England/CAMB-1B1182/2020|EPI_ISL_459166|20</td><td>....................</td><td>....................</td><td>.....................</td><td>....................</td><td>....................</td><td>....................</td><td></td></tr>
<tr><td>C</td><td>3981</td><td>hCoV-19/England/CAMB-1B08E8/2020|EPI_ISL_459167|20</td><td>....................</td><td>....................</td><td>.....................</td><td>....................</td><td>....................</td><td>....................</td><td></td></tr>
<tr><td>C</td><td>3982</td><td>hCoV-19/England/CAMB-1B087F/2020|EPI_ISL_459169|20</td><td>....................</td><td>....................</td><td>.....................</td><td>....................</td><td>....................</td><td>....................</td><td></td></tr>
<tr><td>C</td><td>3983</td><td>hCoV-19/England/CAMB-1AFE62/2020|EPI_ISL_459171|20</td><td>....................</td><td>....................</td><td>.....................</td><td>....................</td><td>....................</td><td>....................</td><td></td></tr>
<tr><td>C</td><td>3984</td><td>hCoV-19/England/CAMB-1B0930/2020|EPI_ISL_459174|20</td><td>....................</td><td>....................</td><td>.....................</td><td>....................</td><td>....................</td><td>....................</td><td></td></tr>
<tr><td>C</td><td>3985</td><td>hCoV-19/England/CAMB-1B1261/2020|EPI_ISL_459178|20</td><td>....................</td><td>....................</td><td>.....................</td><td>....................</td><td>....................</td><td>....................</td><td></td></tr>
<tr><td>C</td><td>3986</td><td>hCoV-19/England/CAMB-1AFB6B/2020|EPI_ISL_459179|20</td><td>....................</td><td>....................</td><td>.....................</td><td>....................</td><td>....................</td><td>....................</td><td></td></tr>
<tr><td>C</td><td>3987</td><td>hCoV-19/England/CAMB-1AFB7A/2020|EPI_ISL_459194|20</td><td>....................</td><td>....................</td><td>.....................</td><td>....................</td><td>....................</td><td>....................</td><td></td></tr>
<tr><td>C</td><td>3988</td><td>hCoV-19/USA/MA-MGH-00687/2020|EPI_ISL_460132|2020-</td><td>....................</td><td>....................</td><td>.....................</td><td>....................</td><td>....................</td><td>....................</td><td></td></tr>
<tr><td>C</td><td>3989</td><td>hCoV-19/USA/MA-MGH-00289/2020|EPI_ISL_460133|2020-</td><td>....................</td><td>....................</td><td>.....................</td><td>....................</td><td>....................</td><td>....................</td><td></td></tr>
<tr><td>C</td><td>3990</td><td>hCoV-19/USA/MA-MGH-00611/2020|EPI_ISL_460134|2020-</td><td>....................</td><td>....................</td><td>.....................</td><td>....................</td><td>....................</td><td>....................</td><td></td></tr>
<tr><td>C</td><td>3991</td><td>hCoV-19/USA/MA-MGH-00661/2020|EPI_ISL_460135|2020-</td><td>....................</td><td>....................</td><td>.....................</td><td>....................</td><td>....................</td><td>....................</td><td></td></tr>
<tr><td>C</td><td>3992</td><td>hCoV-19/USA/MA-MGH-00628/2020|EPI_ISL_460137|2020-</td><td>....................</td><td>....................</td><td>.....................</td><td>....................</td><td>....................</td><td>....................</td><td></td></tr>
<tr><td>C</td><td>3993</td><td>hCoV-19/USA/MA-MGH-00544/2020|EPI_ISL_460138|2020-</td><td>....................</td><td>....................</td><td>.....................</td><td>....................</td><td>....................</td><td>....................</td><td></td></tr>
<tr><td>C</td><td>3994</td><td>hCoV-19/USA/MA-MGH-00651/2020|EPI_ISL_460139|2020-</td><td>....................</td><td>....................</td><td>.....................</td><td>....................</td><td>....................</td><td>....................</td><td></td></tr>
<tr><td>C</td><td>3995</td><td>hCoV-19/USA/MA-MGH-00569/2020|EPI_ISL_460140|2020-</td><td>....................</td><td>....................</td><td>.....................</td><td>....................</td><td>....................</td><td>....................</td><td></td></tr>
<tr><td>C</td><td>3996</td><td>hCoV-19/USA/MA-MGH-00266/2020|EPI_ISL_460141|2020-</td><td>....................</td><td>....................</td><td>.....................</td><td>....................</td><td>....................</td><td>....................</td><td></td></tr>
<tr><td>C</td><td>3997</td><td>hCoV-19/USA/MA-MGH-00210/2020|EPI_ISL_460142|2020-</td><td>....................</td><td>....................</td><td>.....................</td><td>....................</td><td>....................</td><td>....................</td><td></td></tr>
<tr><td>C</td><td>3998</td><td>hCoV-19/USA/MA-MGH-00054/2020|EPI_ISL_460143|2020-</td><td>....................</td><td>....................</td><td>.....................</td><td>....................</td><td>....................</td><td>....................</td><td></td></tr>
<tr><td>C</td><td>3999</td><td>hCoV-19/USA/MA-MGH-00274/2020|EPI_ISL_460144|2020-</td><td>....................</td><td>....................</td><td>.....................</td><td>....................</td><td>....................</td><td>....................</td><td></td></tr>
</table>

| | | | | | | | | |
|---|---|---|---|---|---|---|---|---|
| C | 4000 hCoV-19/USA/MA-MGH-00086/2020\|EPI_ISL_460145\|2020- | ···················· | ···················· | ····················· | ···················· | ···················· | ···················· | |
| C | 4001 hCoV-19/USA/MA-MGH-00683/2020\|EPI_ISL_460146\|2020- | ···················· | ···················· | ····················· | ···················· | ···················· | ···················· | |
| C | 4002 hCoV-19/USA/MA-MGH-00242/2020\|EPI_ISL_460147\|2020- | ···················· | ···················· | ····················· | ···················· | ···················· | ···················· | |
| C | 4003 hCoV-19/USA/MA-MGH-00642/2020\|EPI_ISL_460148\|2020- | ···················· | ···················· | ····················· | ···················· | ···················· | ···················· | |
| C | 4004 hCoV-19/USA/MA-MGH-00023/2020\|EPI_ISL_460149\|2020- | ···················· | ···················· | ····················· | ···················· | ···················· | ···················· | |
| C | 4005 hCoV-19/USA/MA-MGH-00021/2020\|EPI_ISL_460150\|2020- | ···················· | ···················· | ····················· | ···················· | ···················· | ···················· | |
| C | 4006 hCoV-19/USA/MA-MGH-00675/2020\|EPI_ISL_460153\|2020- | ···················· | ······A············· | ····················· | ···················· | ···················· | ···················· | |
| C | 4007 hCoV-19/USA/MA-MGH-00013/2020\|EPI_ISL_460154\|2020- | ···················· | ···················· | ····················· | ···················· | ···················· | ···················· | |
| C | 4008 hCoV-19/USA/MA-MGH-00288/2020\|EPI_ISL_460155\|2020- | ···················· | ···················· | ····················· | ···················· | ···················· | ···················· | |
| C | 4009 hCoV-19/USA/MA-MGH-00502/2020\|EPI_ISL_460156\|2020- | ···················· | ···················· | ····················· | ···················· | ···················· | ···················· | |
| C | 4010 hCoV-19/USA/MA-MGH-00297/2020\|EPI_ISL_460157\|2020- | ···················· | ···················· | ····················· | ···················· | ···················· | ···················· | |
| C | 4011 hCoV-19/USA/MA-MGH-00433/2020\|EPI_ISL_460158\|2020- | ···················· | ···················· | ····················· | ···················· | ···················· | ···················· | |
| C | 4012 hCoV-19/USA/MA-MGH-00467/2020\|EPI_ISL_460159\|2020- | ···················· | ···················· | ····················· | ···················· | ···················· | ···················· | |
| C | 4013 hCoV-19/USA/MA-MGH-00264/2020\|EPI_ISL_460160\|2020- | ···················· | ···················· | ····················· | ···················· | ···················· | ···················· | |
| C | 4014 hCoV-19/USA/MA-MGH-00603/2020\|EPI_ISL_460161\|2020- | ···················· | ···················· | ····················· | ···················· | ···················· | ···················· | |
| C | 4015 hCoV-19/USA/MA-MGH-00691/2020\|EPI_ISL_460162\|2020- | ···················· | ···················· | ····················· | ···················· | ···················· | ···················· | |
| C | 4016 hCoV-19/USA/MA-MGH-00602/2020\|EPI_ISL_460163\|2020- | ···················· | ···················· | ····················· | ···················· | ···················· | ···················· | |
| C | 4017 hCoV-19/USA/MA-MGH-00269/2020\|EPI_ISL_460164\|2020- | ···················· | ···················· | ····················· | ···················· | ···················· | ···················· | |
| C | 4018 hCoV-19/USA/MA-MGH-00666/2020\|EPI_ISL_460165\|2020- | ···················· | ···················· | ····················· | ···················· | ···················· | ···················· | |
| C | 4020 hCoV-19/USA/MA-MGH-00471/2020\|EPI_ISL_460168\|2020- | ···················· | ···················· | ····················· | ···················· | ···················· | ···················· | |
| C | 4021 hCoV-19/USA/MA-MGH-00466/2020\|EPI_ISL_460176\|2020- | ···················· | ···················· | ····················· | ···················· | ···················· | ···················· | |
| C | 4022 hCoV-19/USA/MA-MGH-00464/2020\|EPI_ISL_460177\|2020- | ···················· | ···················· | ····················· | ···················· | ···················· | ···················· | |
| C | 4023 hCoV-19/USA/MA-MGH-00530/2020\|EPI_ISL_460178\|2020- | ···················· | ···················· | ····················· | ···················· | ···················· | ···················· | |
| C | 4024 hCoV-19/USA/MA-MGH-00174/2020\|EPI_ISL_460179\|2020- | ···················· | ···················· | ····················· | ···················· | ···················· | ···················· | |
| C | 4025 hCoV-19/USA/MA-MGH-00570/2020\|EPI_ISL_460180\|2020- | ···················· | ···················· | ····················· | ···················· | ···················· | ···················· | |
| C | 4026 hCoV-19/USA/MA-MGH-00283/2020\|EPI_ISL_460181\|2020- | ···················· | ···················· | ····················· | ···················· | ···················· | ···················· | |
| C | 4027 hCoV-19/USA/MA-MGH-00542/2020\|EPI_ISL_460182\|2020- | ···················· | ···················· | ····················· | ···················· | ···················· | ···················· | |
| C | 4028 hCoV-19/USA/MA-MGH-00161/2020\|EPI_ISL_460183\|2020- | ···················· | ···················· | ····················· | ···················· | ···················· | ···················· | |
| C | 4029 hCoV-19/USA/MA-MGH-00089/2020\|EPI_ISL_460184\|2020- | ···················· | ···················· | ····················· | ···················· | ···················· | ···················· | |
| C | 4030 hCoV-19/USA/MA-MGH-00488/2020\|EPI_ISL_460185\|2020- | ···················· | ···················· | ····················· | ···················· | ···················· | ···················· | |
| C | 4031 hCoV-19/USA/MA-MGH-00563/2020\|EPI_ISL_460186\|2020- | ···················· | ···················· | ····················· | ···················· | ···················· | ···················· | |
| C | 4032 hCoV-19/USA/MA-MGH-00690/2020\|EPI_ISL_460187\|2020- | ···················· | ···················· | ····················· | ···················· | ···················· | ···················· | |
| C | 4033 hCoV-19/USA/MA-MGH-00229/2020\|EPI_ISL_460188\|2020- | ···················· | ···················· | ····················· | ···················· | ···················· | ···················· | |
| C | 4034 hCoV-19/USA/MA-MGH-00633/2020\|EPI_ISL_460189\|2020- | ···················· | ···················· | ····················· | ···················· | ···················· | ···················· | |
| C | 4035 hCoV-19/USA/MA-MGH-00669/2020\|EPI_ISL_460190\|2020- | ···················· | ···················· | ····················· | ···················· | ···················· | ···················· | |
| C | 4036 hCoV-19/USA/MA-MGH-00695/2020\|EPI_ISL_460191\|2020- | ···················· | ···················· | ····················· | ···················· | ···················· | ···················· | |
| C | 4037 hCoV-19/USA/MA-MGH-00281/2020\|EPI_ISL_460195\|2020- | ···················· | ···················· | ····················· | ···················· | ···················· | ···················· | |
| C | 4038 hCoV-19/USA/MA-MGH-00290/2020\|EPI_ISL_460196\|2020- | ···················· | ···················· | ····················· | ···················· | ···················· | ···················· | |
| C | 4039 hCoV-19/USA/MA-MGH-00653/2020\|EPI_ISL_460197\|2020- | ···················· | ···················· | ····················· | ···················· | ···················· | ···················· | |
| C | 4040 hCoV-19/USA/MA-MGH-00445/2020\|EPI_ISL_460198\|2020- | ···················· | ···················· | ····················· | ···················· | ···················· | ···················· | |
| C | 4041 hCoV-19/USA/WA-UW-4698/2020\|EPI_ISL_461402\|2020-04 | ···················· | ···················· | ····················· | ···················· | ···················· | ···················· | |
| C | 4042 hCoV-19/USA/WA-UW-4765/2020\|EPI_ISL_461403\|2020-03 | ···················· | ···················· | ····················· | ···················· | ···················· | ···················· | |
| C | 4043 hCoV-19/USA/WA-UW-4917/2020\|EPI_ISL_461404\|2020-04 | ···················· | ···················· | ····················· | ···················· | ···················· | ···················· | |
| C | 4044 hCoV-19/USA/WA-UW-4931/2020\|EPI_ISL_461405\|2020-04 | ···················· | ···················· | ····················· | ···················· | ···················· | ···················· | |
| C | 4045 hCoV-19/USA/WA-UW-8437/2020\|EPI_ISL_461406\|2020-04 | ···················· | ···················· | ····················· | ···················· | ···················· | ···················· | |
| C | 4046 hCoV-19/USA/WA-UW-8527/2020\|EPI_ISL_461407\|2020-05 | ···················· | ···················· | ····················· | ···················· | ···················· | ···················· | |
| C | 4047 hCoV-19/USA/CT-UW-6577/2020\|EPI_ISL_461408\|2020-04 | ···················· | ···················· | ····················· | ···················· | ···················· | ···················· | |
| C | 4048 hCoV-19/USA/CT-UW-6582/2020\|EPI_ISL_461409\|2020-04 | ···················· | ···················· | ····················· | ···················· | ···················· | ···················· | |
| C | 4049 hCoV-19/USA/WA-UW-6585/2020\|EPI_ISL_461410\|2020-04 | ···················· | ···················· | ····················· | ···················· | ···················· | ···················· | |
| C | 4050 hCoV-19/USA/CT-UW-6586/2020\|EPI_ISL_461411\|2020-04 | ···················· | ···················· | ····················· | ···················· | ···················· | ···················· | |
| C | 4051 hCoV-19/USA/CT-UW-6587/2020\|EPI_ISL_461412\|2020-04 | ···················· | ···················· | ····················· | ···················· | ···················· | ···················· | |
| C | 4052 hCoV-19/USA/WA-UW-6589/2020\|EPI_ISL_461413\|2020-04 | ···················· | ···················· | ····················· | ···················· | ···················· | ···················· | |
| C | 4053 hCoV-19/USA/CT-UW-6590/2020\|EPI_ISL_461414\|2020-04 | ···················· | ···················· | ····················· | ···················· | ···················· | ···················· | |
| C | 4054 hCoV-19/USA/WA-UW-6591/2020\|EPI_ISL_461415\|2020-04 | ···················· | ···················· | ····················· | ···················· | ···················· | ···················· | |
| C | 4055 hCoV-19/USA/CT-UW-6592/2020\|EPI_ISL_461416\|2020-04 | ···················· | ···················· | ····················· | ···················· | ···················· | ···················· | |
| C | 4056 hCoV-19/USA/CT-UW-6594/2020\|EPI_ISL_461417\|2020-04 | ···················· | ···················A | ····················· | ···················· | ···················· | ···················· | 1 |
| C | 4057 hCoV-19/USA/WA-UW-6596/2020\|EPI_ISL_461418\|2020-04 | ···················· | ···················· | ····················· | ···················· | ···················· | ···················· | |
| C | 4058 hCoV-19/USA/WA-UW-6601/2020\|EPI_ISL_461419\|2020-04 | ···················· | ···················· | ····················· | ···················· | ···················· | ···················· | |
| C | 4059 hCoV-19/USA/WA-UW-10091/2020\|EPI_ISL_461461\|2020-0 | ···················· | ···················· | ····················· | ···················· | ···················· | ···················· | |
| C | 4060 hCoV-19/USA/WA-UW-10093/2020\|EPI_ISL_461462\|2020-0 | ···················· | ···················· | ····················· | ···················· | ···················· | ···················· | |
| C | 4061 hCoV-19/USA/WA-UW-10094/2020\|EPI_ISL_461463\|2020-0 | ···················· | ···················· | ····················· | ···················· | ···················· | ···················· | |
| C | 4062 hCoV-19/USA/WA-UW-10102/2020\|EPI_ISL_461464\|2020-0 | ···················· | ···················· | ····················· | ···················· | ···················· | ···················· | |
| C | 4063 hCoV-19/USA/WA-UW-10106/2020\|EPI_ISL_461465\|2020-0 | ···················· | ···················· | ····················· | ···················· | ···················· | ···················· | |
| C | 4064 hCoV-19/USA/WA-UW-10108/2020\|EPI_ISL_461467\|2020-0 | ···················· | ···················· | ····················· | ···················· | ···················· | ···················· | |
| C | 4065 hCoV-19/USA/WA-UW-10114/2020\|EPI_ISL_461468\|2020-0 | ···················· | ···················· | ····················· | ···················· | ···················· | ···················· | |
| C | 4066 hCoV-19/USA/WA-UW-10117/2020\|EPI_ISL_461469\|2020-0 | ···················· | ···················· | ····················· | ···················· | ···················· | ···················· | |
| C | 4067 hCoV-19/USA/WA-UW-10118/2020\|EPI_ISL_461470\|2020-0 | ···················· | ···················· | ····················· | ···················· | ···················· | ···················· | |
| C | 4068 hCoV-19/USA/WA-UW-10127/2020\|EPI_ISL_461471\|2020-0 | ···················· | ···················· | ····················· | ···················· | ···················· | ···················· | |
| C | 4069 hCoV-19/USA/WA-UW-10128/2020\|EPI_ISL_461472\|2020-0 | ···················· | ···················· | ····················· | ···················· | ···················· | ···················· | |
| C | 4070 hCoV-19/USA/WA-UW-10129/2020\|EPI_ISL_461473\|2020-0 | ···················· | ···················· | ····················· | ···················· | ···················· | ···················· | |
| C | 4071 hCoV-19/USA/WA-UW-10130/2020\|EPI_ISL_461474\|2020-0 | ···················· | ···················· | ····················· | ···················· | ···················· | ···················· | |
| C | 4072 hCoV-19/USA/WA-UW-10131/2020\|EPI_ISL_461475\|2020-0 | ···················· | ···················· | ····················· | ···················· | ···················· | ···················· | |
| C | 4073 hCoV-19/USA/WA-UW-10136/2020\|EPI_ISL_461476\|2020-0 | ···················· | ···················· | ····················· | ···················· | ···················· | ···················· | |
| C | 4074 hCoV-19/USA/WA-UW-10138/2020\|EPI_ISL_461477\|2020-0 | ···················· | ···················· | ····················· | ···················· | ···················· | ···················· | |
| C | 4075 hCoV-19/India/GJ-GBRC151a/2020\|EPI_ISL_461478\|2020 | ···················· | ···················· | ····················· | ···················· | ···················· | ···················· | |
| C | 4076 hCoV-19/India/GJ-GBRC151b/2020\|EPI_ISL_461479\|2020 | ···················· | ···················· | ····················· | ···················· | ···················· | ···················· | |
| C | 4077 hCoV-19/India/GJ-GBRC152/2020\|EPI_ISL_461480\|2020- | ···················· | ···················· | ····················· | ···················· | ···················· | ···················· | |
| C | 4078 hCoV-19/India/GJ-GBRC153a/2020\|EPI_ISL_461481\|2020 | ···················· | ···················· | ····················· | ···················· | ···················· | ···················· | |

| | | | | | | | |
|---|---|---|---|---|---|---|---|
| C | 4079 hCoV-19/India/GJ-GBRC153b/2020\|EPI_ISL_461482\|2020 | ···················· | ···················· | ····················· | ···················· | ···················· | ···················· |
| C | 4080 hCoV-19/India/GJ-GBRC154/2020\|EPI_ISL_461483\|2020- | ···················· | ···················· | ····················· | ···················· | ···················· | ···················· |
| C | 4081 hCoV-19/India/GJ-GBRC155/2020\|EPI_ISL_461484\|2020- | ···················· | ·A·················· | ····················· | ···················· | ···················· | ···A················ |
| C | 4082 hCoV-19/India/GJ-GBRC156/2020\|EPI_ISL_461485\|2020- | ···················· | ·A·················· | ····················· | ···················· | ···················· | ···A················ |
| C | 4083 hCoV-19/India/GJ-GBRC157a/2020\|EPI_ISL_461486\|2020 | ···················· | ···················· | ····················· | ···················· | ···················· | ···················· |
| C | 4084 hCoV-19/India/GJ-GBRC157b/2020\|EPI_ISL_461487\|2020 | ···················· | ···················· | ····················· | ···················· | ···················· | ···················· |
| C | 4085 hCoV-19/India/GJ-GBRC158/2020\|EPI_ISL_461488\|2020- | ···················· | ·A·················· | ····················· | ···················· | ···················· | ···A················ |
| C | 4086 hCoV-19/India/GJ-GBRC159a/2020\|EPI_ISL_461489\|2020 | ···················· | ·A·················· | ····················· | ···················· | ···················· | ···A················ |
| C | 4087 hCoV-19/India/GJ-GBRC159b/2020\|EPI_ISL_461490\|2020 | ···················· | ···················· | ····················· | ···················· | ···················· | ···A················ |
| C | 4088 hCoV-19/India/GJ-GBRC160a/2020\|EPI_ISL_461491\|2020 | ···················· | ·A·················· | ····················· | ···················· | ···················· | ···A················ |
| C | 4089 hCoV-19/India/GJ-GBRC160b/2020\|EPI_ISL_461492\|2020 | ···················· | ···················· | ····················· | ···················· | ···················· | ···················· |
| C | 4090 hCoV-19/India/GJ-GBRC161a/2020\|EPI_ISL_461493\|2020 | ···················· | ·A·················· | ····················· | ···················· | ···················· | ···················· |
| C | 4091 hCoV-19/India/GJ-GBRC161b/2020\|EPI_ISL_461494\|2020 | ···················· | ···················· | ····················· | ···················· | ···················· | ···················· |
| C | 4092 hCoV-19/India/GJ-GBRC162a/2020\|EPI_ISL_461495\|2020 | ···················· | ·A·················· | ····················· | ···················· | ···················· | ···A················ |
| C | 4093 hCoV-19/India/GJ-GBRC162b/2020\|EPI_ISL_461496\|2020 | ···················· | ·A·················· | ····················· | ···················· | ···················· | ···················· |
| C | 4094 hCoV-19/India/GJ-GBRC163a/2020\|EPI_ISL_461497\|2020 | ···················· | ···················· | ····················· | ···················· | ···················· | ···················· |
| C | 4095 hCoV-19/India/GJ-GBRC163b/2020\|EPI_ISL_461498\|2020 | ···················· | ···················· | ····················· | ···················· | ···················· | ···················· |
| C | 4096 hCoV-19/India/GJ-GBRC164/2020\|EPI_ISL_461499\|2020- | ···················· | ·A·················· | ····················· | ···················· | ···················· | ···A················ |
| C | 4097 hCoV-19/England/NOTT-110C80/2020\|EPI_ISL_448400\|20 | ···················· | ···················· | ····················· | ···················· | ···················· | ···················· |
| C | 4098 hCoV-19/England/NOTT-110C9F/2020\|EPI_ISL_448401\|20 | ···················· | ···················· | ····················· | ···················· | ···················· | ···················· |
| C | 4099 hCoV-19/England/NOTT-110CAE/2020\|EPI_ISL_448402\|20 | ···················· | ···················· | ····················· | ···················· | ···················· | ···················· |
| C | 4100 hCoV-19/England/NOTT-110CCC/2020\|EPI_ISL_448404\|20 | ···················· | ···················· | ····················· | ···················· | ···················· | ···················· |
| C | 4101 hCoV-19/England/NOTT-110CDB/2020\|EPI_ISL_448405\|20 | ···················· | ···················· | ····················· | ···················· | ···················· | ···················· |
| C | 4102 hCoV-19/England/NOTT-110CEA/2020\|EPI_ISL_448406\|20 | ···················· | ···················· | ····················· | ···················· | ···················· | ···················· |
| C | 4104 hCoV-19/England/NOTT-110D05/2020\|EPI_ISL_448408\|20 | ···················· | ···················· | ····················· | ···················· | ···················· | ···················· |
| C | 4105 hCoV-19/England/NOTT-110D14/2020\|EPI_ISL_448409\|20 | ···················· | ···················· | ····················· | ···················· | ···················· | ···················· |
| C | 4106 hCoV-19/England/NOTT-110D23/2020\|EPI_ISL_448410\|20 | ···················· | ···················· | ····················· | ···················· | ···················· | ···················· |
| C | 4107 hCoV-19/England/NOTT-110D41/2020\|EPI_ISL_448412\|20 | ···················· | ···················· | ····················· | ···················· | ···················· | ···················· |
| C | 4108 hCoV-19/England/NOTT-110D50/2020\|EPI_ISL_448413\|20 | ···················· | ···················· | ····················· | ···················· | ···················· | ···················· |
| C | 4109 hCoV-19/England/NOTT-110D6F/2020\|EPI_ISL_448414\|20 | ···················· | ···················· | ····················· | ···················· | ···················· | ···················· |
| C | 4110 hCoV-19/England/NOTT-110D7E/2020\|EPI_ISL_448415\|20 | ···················· | ···················· | ····················· | ···················· | ···················· | ···················· |
| C | 4111 hCoV-19/England/NOTT-110D8D/2020\|EPI_ISL_448416\|20 | ···················· | ···················· | ····················· | ···················· | ···················· | ···················· |
| C | 4112 hCoV-19/England/OXON-AC552/2020\|EPI_ISL_448460\|202 | ···················· | ···················· | ····················· | ···················· | ···················· | ···················· |
| C | 4113 hCoV-19/England/OXON-AC561/2020\|EPI_ISL_448461\|202 | ···················· | ·············A······ | ····················· | ···················· | ···················· | ···················· |
| C | 4114 hCoV-19/England/OXON-AC570/2020\|EPI_ISL_448462\|202 | ···················· | ···················· | ····················· | ···················· | ···················· | ···················· |
| C | 4115 hCoV-19/England/OXON-AC5AD/2020\|EPI_ISL_448463\|202 | ···················· | ···················· | ····················· | ···················· | ···················· | ···················· |
| C | 4116 hCoV-19/England/OXON-AC5CB/2020\|EPI_ISL_448465\|202 | ···················· | ···················· | ····················· | ···················· | ···················· | ···················· |
| C | 4117 hCoV-19/England/OXON-AC604/2020\|EPI_ISL_448467\|202 | ···················· | ···················· | ····················· | ···················· | ···················· | ···················· |
| C | 4118 hCoV-19/England/OXON-AC613/2020\|EPI_ISL_448468\|202 | ···················· | ···················· | ····················· | ···················· | ···················· | ···················· |
| C | 4119 hCoV-19/England/OXON-AC622/2020\|EPI_ISL_448469\|202 | ···················· | ···················· | ····················· | ···················· | ···················· | ···················· |
| C | 4120 hCoV-19/England/OXON-AC631/2020\|EPI_ISL_448470\|202 | ···················· | ···················· | ····················· | ···················· | ···················· | ···················· |
| C | 4121 hCoV-19/England/OXON-AC640/2020\|EPI_ISL_448471\|202 | ···················· | ···················· | ····················· | ···················· | ···················· | ···················· |
| C | 4122 hCoV-19/England/OXON-AC67D/2020\|EPI_ISL_448472\|202 | ···················· | ···················· | ····················· | ···················· | ···················· | ···················· |
| C | 4123 hCoV-19/England/OXON-AC68C/2020\|EPI_ISL_448473\|202 | ···················· | ···················· | ····················· | ···················· | ···················· | ···················· |
| C | 4124 hCoV-19/England/OXON-AC6D7/2020\|EPI_ISL_448476\|202 | ···················· | ···················· | ····················· | ···················· | ···················· | ···················· |
| C | 4125 hCoV-19/England/OXON-AC6F5/2020\|EPI_ISL_448477\|202 | ···················· | ···················· | ····················· | ···················· | ···················· | ···················· |
| C | 4126 hCoV-19/England/OXON-AC710/2020\|EPI_ISL_448478\|202 | ···················· | ···················· | ····················· | ···················· | ···················· | ···················· |
| C | 4127 hCoV-19/England/OXON-AC72F/2020\|EPI_ISL_448479\|202 | ···················· | ···················· | ····················· | ···················· | ···················· | ···················· |
| C | 4128 hCoV-19/England/OXON-AC73E/2020\|EPI_ISL_448480\|202 | ···················· | ···················· | ····················· | ···················· | ···················· | ···················· |
| C | 4129 hCoV-19/England/OXON-AC74D/2020\|EPI_ISL_448481\|202 | ···················· | ···················· | ····················· | ···················· | ···················· | ···················· |
| C | 4130 hCoV-19/England/OXON-AC789/2020\|EPI_ISL_448482\|202 | ···················· | ···················· | ····················· | ···················· | ···················· | ···················· |
| C | 4131 hCoV-19/England/OXON-AC7F2/2020\|EPI_ISL_448485\|202 | ···················· | ···················· | ····················· | ···················· | ···················· | ···················· |
| C | 4132 hCoV-19/England/OXON-AC895/2020\|EPI_ISL_448490\|202 | ···················· | ···················· | ····················· | ···················· | ···················· | ···················· |
| C | 4133 hCoV-19/England/OXON-AC8D1/2020\|EPI_ISL_448491\|202 | ···················· | ···················· | ····················· | ···················· | ···················· | ···················· |
| C | 4134 hCoV-19/England/OXON-AC8FF/2020\|EPI_ISL_448492\|202 | ···················· | ···················· | ····················· | ···················· | ···················· | ···················· |
| C | 4135 hCoV-19/England/OXON-AC90B/2020\|EPI_ISL_448493\|202 | ···················· | ···················· | ····················· | ···················· | ···················· | ···················· |
| C | 4136 hCoV-19/England/OXON-AC91A/2020\|EPI_ISL_448494\|202 | ···················· | ···················· | ····················· | ···················· | ···················· | ···················· |
| C | 4137 hCoV-19/England/OXON-AC929/2020\|EPI_ISL_448495\|202 | ···················· | ···················· | ····················· | ···················· | ···················· | ···················· |
| C | 4138 hCoV-19/England/OXON-AC938/2020\|EPI_ISL_448496\|202 | ···················· | ···················· | ····················· | ···················· | ···················· | ···················· |
| C | 4139 hCoV-19/England/OXON-AC965/2020\|EPI_ISL_448497\|202 | ···················· | ···················· | ····················· | ···················· | ···················· | ···················· |
| C | 4140 hCoV-19/England/OXON-AC983/2020\|EPI_ISL_448498\|202 | ···················· | ···················· | ····················· | ···················· | ···················· | ···················· |
| C | 4141 hCoV-19/England/OXON-AC9A1/2020\|EPI_ISL_448499\|202 | ···················· | ···················· | ····················· | ···················· | ···················· | ···················· |
| C | 4142 hCoV-19/England/LOND-D55D8/2020\|EPI_ISL_449702\|202 | ···················· | ···················· | ····················· | ···················· | ···················· | ···················· |
| C | 4143 hCoV-19/England/LOND-D55E7/2020\|EPI_ISL_449703\|202 | ···················· | ···················· | ····················· | ···················· | ···················· | ···················· |
| C | 4144 hCoV-19/England/LOND-D567B/2020\|EPI_ISL_449710\|202 | ···················· | ···················· | ····················· | ···················· | ···················· | ···················· |
| C | 4145 hCoV-19/England/LOND-D568A/2020\|EPI_ISL_449711\|202 | ···················· | ···················· | ····················· | ···················· | ···················· | ···················· |
| C | 4146 hCoV-19/England/LOND-D5699/2020\|EPI_ISL_449712\|202 | ···················· | ···················· | ····················· | ···················· | ···················· | ···················· |
| C | 4147 hCoV-19/England/LOND-D56A8/2020\|EPI_ISL_449713\|202 | ···················· | ···················· | ····················· | ···················· | ···················· | ···················· |
| C | 4148 hCoV-19/England/LOND-D56C6/2020\|EPI_ISL_449714\|202 | ···················· | ···················· | ····················· | ···················· | ···················· | ···················· |
| C | 4149 hCoV-19/England/LOND-D580C/2020\|EPI_ISL_449726\|202 | ···················· | ···················· | ····················· | ···················· | ···················· | ···················· |
| C | 4150 hCoV-19/England/LOND-D582A/2020\|EPI_ISL_449727\|202 | ···················· | ···················· | ····················· | ···················· | ···················· | ···················· |
| C | 4151 hCoV-19/England/LOND-D5839/2020\|EPI_ISL_449728\|202 | ···················· | ···················· | ····················· | ···················· | ···················· | ···················· |
| C | 4152 hCoV-19/England/LOND-D5848/2020\|EPI_ISL_449729\|202 | ···················· | ···················· | ····················· | ···················· | ···················· | ···················· |
| C | 4153 hCoV-19/Norway/2386/2020\|EPI_ISL_449791\|2020-04-04 | ···················· | ···················· | ····················· | ···················· | ···················· | ···················· |
| C | 4154 hCoV-19/Norway/2387/2020\|EPI_ISL_449792\|2020-04-05 | ···················· | ···················· | ····················· | ···················· | ···················· | ···················· |
| C | 4155 hCoV-19/Norway/2388/2020\|EPI_ISL_449793\|2020-04-02 | ···················· | ···················· | ····················· | ···················· | ···················· | ···················· |
| C | 4156 hCoV-19/Norway/2390/2020\|EPI_ISL_449794\|2020-04-02 | ···················· | ···················· | ····················· | ···················· | ···················· | ···················· |
| C | 4157 hCoV-19/Slovenia/MB0419/2020\|EPI_ISL_449799\|2020-0 | ···················· | ···················· | ····················· | ···················· | ···················· | ···················· |

| | | | | | | | | |
|---|---|---|---|---|---|---|---|---|
| C | 4158 | hCoV-19/USA/WI-UW-20/2020\|EPI_ISL_450700\|2020-03-1 | ···················· | ···················· | ···················· | ···················· | ···················· | ···················· |
| C | 4159 | hCoV-19/USA/IL-UW-347/2020\|EPI_ISL_450701\|2020-04- | ···················· | ···················· | ···················· | ···················· | ···················· | ···················· |
| C | 4160 | hCoV-19/USA/WI-UW-356/2020\|EPI_ISL_450710\|2020-04- | ···················· | ···················· | ···················· | ···················· | ···················· | ···················· |
| C | 4161 | hCoV-19/USA/IL-UW-357/2020\|EPI_ISL_450711\|2020-04- | ···················· | ···················· | ···················· | ···················· | ···················· | ···················· |
| C | 4162 | hCoV-19/USA/IL-UW-358/2020\|EPI_ISL_450712\|2020-04- | ···················· | ···················· | ···················· | ···················· | ···················· | ···················· |
| C | 4163 | hCoV-19/USA/WI-UW-359/2020\|EPI_ISL_450713\|2020-04- | ···················· | ···················· | ···················· | ···················· | ···················· | ···················· |
| C | 4164 | hCoV-19/USA/IL-UW-360/2020\|EPI_ISL_450714\|2020-04- | ···················· | ···················· | ···················· | ···················· | ···················· | ···················· |
| C | 4165 | hCoV-19/USA/IL-UW-361/2020\|EPI_ISL_450715\|2020-04- | ···················· | ···················· | ···················· | ···················· | ···················· | ···················· |
| C | 4166 | hCoV-19/USA/WI-UW-362/2020\|EPI_ISL_450716\|2020-04- | ···················· | ···················· | ···················· | ···················· | ···················· | ···················· |
| C | 4167 | hCoV-19/USA/IL-UW-363/2020\|EPI_ISL_450717\|2020-04- | ···················· | ···················· | ···················· | ···················· | ···················· | ···················· |
| C | 4168 | hCoV-19/USA/IL-UW-364/2020\|EPI_ISL_450718\|2020-04- | ···················· | ···················· | ···················· | ···················· | ···················· | ···················· |
| C | 4169 | hCoV-19/USA/IL-UW-365/2020\|EPI_ISL_450719\|2020-04- | ···················· | ···················· | ···················· | ···················· | ···················· | ···················· |
| C | 4170 | hCoV-19/USA/IL-UW-366/2020\|EPI_ISL_450720\|2020-04- | ···················· | ···················· | ···················· | ···················· | ···················· | ···················· |
| C | 4171 | hCoV-19/USA/WI-UW-367/2020\|EPI_ISL_450721\|2020-04- | ···················· | ···················· | ···················· | ···················· | ···················· | ···················· |
| C | 4172 | hCoV-19/USA/IL-UW-368/2020\|EPI_ISL_450722\|2020-04- | ···················· | ···················· | ···················· | ···················· | ···················· | ···················· |
| C | 4173 | hCoV-19/Thailand/Bangkok-CONI-0018/2020\|EPI_ISL_45 | ···················· | ···················· | ···················· | ···················· | ···················· | ···················· |
| C | 4174 | hCoV-19/Belgium/ITM_C026/2020\|EPI_ISL_450724\|2020- | ···················· | ···················· | ···················· | ···················· | ···················· | ···················· |
| C | 4175 | hCoV-19/Belgium/ITM_C067/2020\|EPI_ISL_450725\|2020- | ···················· | ···················· | ···················· | ···················· | ···················· | ···················· |
| C | 4176 | hCoV-19/Belgium/ITM_C068/2020\|EPI_ISL_450726\|2020- | ···················· | ···················· | ···················· | ···················· | ···················· | ···················· |
| C | 4177 | hCoV-19/Belgium/ITM_C152/2020\|EPI_ISL_450727\|2020- | ···················· | ···················· | ···················· | ···················· | ···················· | ···················· |
| C | 4178 | hCoV-19/Belgium/ITM_C161/2020\|EPI_ISL_450728\|2020- | ···················· | ···················· | ···················· | ···················· | ···················· | ···················· |
| C | 4179 | hCoV-19/Belgium/ITM_C169/2020\|EPI_ISL_450729\|2020- | ···················· | ···················· | ···················· | ···················· | ···················· | ···················· |
| C | 4180 | hCoV-19/USA/MN-MDH-310/2020\|EPI_ISL_450771\|2020-03 | ···················· | ···················· | ···················· | ···················· | ···················· | ···················· |
| C | 4181 | hCoV-19/USA/MN-MDH-312/2020\|EPI_ISL_450772\|2020-03 | ···················· | ···················· | ···················· | ···················· | ···················· | ···················· |
| C | 4182 | hCoV-19/USA/MN-MDH-313/2020\|EPI_ISL_450773\|2020-03 | ···················· | ···················· | ···················· | ···················· | ···················· | ···················· |
| C | 4183 | hCoV-19/USA/MN-MDH-316/2020\|EPI_ISL_450774\|2020-03 | ···················· | ···················· | ···················· | ···················· | ···················· | ···················· |
| C | 4184 | hCoV-19/USA/MN-MDH-317/2020\|EPI_ISL_450775\|2020-03 | ···················· | ···················· | ···················· | ···················· | ···················· | ···················· |
| C | 4185 | hCoV-19/USA/MN-MDH-318/2020\|EPI_ISL_450776\|2020-03 | ···················· | ···················· | ···················· | ···················· | ···················· | ···················· |
| C | 4186 | hCoV-19/USA/MN-MDH-319/2020\|EPI_ISL_450777\|2020-03 | ···················· | ···················· | ···················· | ···················· | ···················· | ···················· |
| C | 4187 | hCoV-19/USA/MN-MDH-320/2020\|EPI_ISL_450778\|2020-03 | ···················· | ···················· | ···················· | ···················· | ···················· | ···················· |
| C | 4188 | hCoV-19/USA/MN-MDH-321/2020\|EPI_ISL_450779\|2020-03 | ···················· | ···················· | ···················· | ···················· | ···················· | ···················· |
| C | 4189 | hCoV-19/USA/MN-MDH-322/2020\|EPI_ISL_450780\|2020-03 | ···················· | ···················· | ···················· | ···················· | ···················· | ···················· |
| C | 4190 | hCoV-19/India/GJ-GBRC94/2020\|EPI_ISL_450781\|2020-0 | ···················· | ···················· | ···················· | ···················· | ···················· | ···················· |
| C | 4191 | hCoV-19/India/GJ-GBRC95a/2020\|EPI_ISL_450782\|2020- | ···················· | ···················· | ···················· | ···················· | ···················· | ···················· |
| C | 4192 | hCoV-19/India/GJ-GBRC95b/2020\|EPI_ISL_450783\|2020- | ···················· | ·A·················· | ···················· | ···················· | ···················· | ··A················· |
| C | 4193 | hCoV-19/India/GJ-GBRC96/2020\|EPI_ISL_450784\|2020-0 | ···················· | ···················· | ···················· | ···················· | ···················· | ···················· |
| C | 4194 | hCoV-19/India/GJ-GBRC97a/2020\|EPI_ISL_450785\|2020- | ···················· | ···················· | ···················· | ···················· | ···················· | ···················· |
| C | 4195 | hCoV-19/India/GJ-GBRC97b/2020\|EPI_ISL_450786\|2020- | ···················· | ···················· | ···················· | ···················· | ···················· | ···················· |
| C | 4196 | hCoV-19/India/GJ-GBRC98/2020\|EPI_ISL_450787\|2020-0 | ···················· | ···················· | ···················· | ···················· | ···················· | ···················· |
| C | 4197 | hCoV-19/India/GJ-GBRC99/2020\|EPI_ISL_450788\|2020-0 | ···················· | ···················· | ···················· | ···················· | ···················· | ···················· |
| C | 4198 | hCoV-19/India/GJ-GBRC100/2020\|EPI_ISL_450789\|2020- | ···················· | ···················· | ···················· | ···················· | ···················· | ···················· |
| C | 4199 | hCoV-19/India/GJ-GBRC101/2020\|EPI_ISL_450790\|2020- | ···················· | ···················· | ···················· | ···················· | ···················· | ···················· |
| C | 4200 | hCoV-19/India/GJ-GBRC102/2020\|EPI_ISL_450791\|2020- | ···················· | ···················· | ···················· | ···················· | ···················· | ···················· |
| C | 4201 | hCoV-19/Jamaica/JM-CDC-0078/2020\|EPI_ISL_450792\|20 | ···················· | ···················· | ···················· | ···················· | ···················· | ···················· |
| C | 4202 | hCoV-19/Jamaica/JM-CDC-0869/2020\|EPI_ISL_450793\|20 | ···················· | ···················· | ···················· | ···················· | ···················· | ···················· |
| C | 4203 | hCoV-19/Jamaica/JM-CDC-4376/2020\|EPI_ISL_450794\|20 | ···················· | ···················· | ···················· | ···················· | ···················· | ···················· |
| C | 4204 | hCoV-19/Jamaica/JM-CDC-5836/2020\|EPI_ISL_450795\|20 | ···················· | ···················· | ···················· | ···················· | ···················· | ···················· |
| C | 4205 | hCoV-19/Jamaica/JM-CDC-6365/2020\|EPI_ISL_450796\|20 | ···················· | ···················· | ···················· | ···················· | ···················· | ···················· |
| C | 4206 | hCoV-19/Jamaica/JM-CDC-6603/2020\|EPI_ISL_450797\|20 | ···················· | ···················· | ···················· | ···················· | ···················· | ···················· |
| C | 4207 | hCoV-19/Jamaica/JM-CDC-6729/2020\|EPI_ISL_450798\|20 | ···················· | ···················· | ···················· | ···················· | ···················· | ···················· |
| C | 4208 | hCoV-19/Jamaica/JM-CDC-7286/2020\|EPI_ISL_450799\|20 | ···················· | ···················· | ···················· | ···················· | ···················· | ···················· |
| C | 4209 | hCoV-19/Portugal/PT0534/2020\|EPI_ISL_454258\|2020-0 | ···················· | ···················· | ···················· | ···················· | ···················· | ···················· |
| C | 4210 | hCoV-19/Portugal/PT0536/2020\|EPI_ISL_454260\|2020-0 | ···················· | ···················· | ···················· | ···················· | ···················· | ···················· |
| C | 4211 | hCoV-19/Portugal/PT0537/2020\|EPI_ISL_454261\|2020-0 | ···················· | ···················· | ···················· | ···················· | ···················· | ···················· |
| C | 4212 | hCoV-19/Portugal/PT0538/2020\|EPI_ISL_454262\|2020-0 | ···················· | ···················· | ···················· | ···················· | ···················· | ···················· |
| C | 4213 | hCoV-19/Portugal/PT0539/2020\|EPI_ISL_454263\|2020-0 | ···················· | ···················· | ···················· | ···················· | ···················· | ···················· |
| C | 4214 | hCoV-19/Portugal/PT0540/2020\|EPI_ISL_454264\|2020-0 | ···················· | ···················· | ···················· | ···················· | ···················· | ···················· |
| C | 4215 | hCoV-19/Portugal/PT0542/2020\|EPI_ISL_454266\|2020-0 | ···················· | ···················· | ···················· | ···················· | ···················· | ···················· |
| C | 4216 | hCoV-19/Portugal/PT0543/2020\|EPI_ISL_454267\|2020-0 | ···················· | ···················· | ···················· | ···················· | ···················· | ···················· |
| C | 4217 | hCoV-19/Portugal/PT0544/2020\|EPI_ISL_454268\|2020-0 | ···················· | ···················· | ···················· | ···················· | ···················· | ···················· |
| C | 4218 | hCoV-19/Portugal/PT0545/2020\|EPI_ISL_454269\|2020-0 | ···················· | ···················· | ···················· | ···················· | ···················· | ···················· |
| C | 4219 | hCoV-19/Portugal/PT0546/2020\|EPI_ISL_454270\|2020-0 | ···················· | ···················· | ···················· | ···················· | ···················· | ···················· |
| C | 4220 | hCoV-19/Portugal/PT0547/2020\|EPI_ISL_454271\|2020-0 | ···················· | ···················· | ···················· | ···················· | ···················· | ···················· |
| C | 4221 | hCoV-19/Portugal/PT0548/2020\|EPI_ISL_454272\|2020-0 | ···················· | ···················· | ···················· | ···················· | ···················· | ···················· |
| C | 4222 | hCoV-19/Portugal/PT0551/2020\|EPI_ISL_454274\|2020-0 | ···················· | ···················· | ···················· | ···················· | ···················· | ···················· |
| C | 4223 | hCoV-19/Portugal/PT0552/2020\|EPI_ISL_454275\|2020-0 | ···················· | ···················· | ···················· | ···················· | ···················· | ···················· |
| C | 4224 | hCoV-19/Portugal/PT0553/2020\|EPI_ISL_454276\|2020-0 | ···················· | ···················· | ···················· | ···················· | ···················· | ···················· |
| C | 4225 | hCoV-19/Portugal/PT0554/2020\|EPI_ISL_454277\|2020-0 | ···················· | ···················· | ···················· | ···················· | ···················· | ···················· |
| C | 4226 | hCoV-19/Portugal/PT0555/2020\|EPI_ISL_454278\|2020-0 | ···················· | ···················· | ···················· | ···················· | ···················· | ···················· |
| C | 4227 | hCoV-19/Portugal/PT0556/2020\|EPI_ISL_454279\|2020-0 | ···················· | ···················· | ···················· | ···················· | ···················· | ···················· |
| C | 4228 | hCoV-19/Portugal/PT0557/2020\|EPI_ISL_454280\|2020-0 | ···················· | ···················· | ···················· | ···················· | ···················· | ···················· |
| C | 4229 | hCoV-19/Portugal/PT0558/2020\|EPI_ISL_454281\|2020-0 | ···················· | ···················· | ···················· | ···················· | ···················· | ···················· |
| C | 4230 | hCoV-19/Portugal/PT0559/2020\|EPI_ISL_454282\|2020-0 | ···················· | ···················· | ···················· | ···················· | ···················· | ···················· |
| C | 4231 | hCoV-19/Portugal/PT0560/2020\|EPI_ISL_454283\|2020-0 | ···················· | ···················· | ···················· | ···················· | ···················· | ···················· |
| C | 4232 | hCoV-19/Portugal/PT0561/2020\|EPI_ISL_454284\|2020-0 | ···················· | ······A············· | ···················· | ···················· | ···················· | ···················· |
| C | 4233 | hCoV-19/Portugal/PT0562/2020\|EPI_ISL_454285\|2020-0 | ···················· | ···················· | ···················· | ···················· | ···················· | ···················· |
| C | 4234 | hCoV-19/Portugal/PT0564/2020\|EPI_ISL_454287\|2020-0 | ···················· | ···················· | ···················· | ···················· | ···················· | ···················· |
| C | 4235 | hCoV-19/Portugal/PT0565/2020\|EPI_ISL_454288\|2020-0 | ···················· | ···················· | ···················· | ···················· | ···················· | ···················· |

| | | | | | | | | |
|---|---|---|---|---|---|---|---|---|
| C | 4236 | hCoV-19/Portugal/PT0566/2020\|EPI_ISL_454289\|2020-0 | ···················· | ···················· | ···················· | ···················· | ···················· | ···················· |
| C | 4237 | hCoV-19/Portugal/PT0567/2020\|EPI_ISL_454290\|2020-0 | ···················· | ···················· | ···················· | ···················· | ···················· | ···················· |
| C | 4238 | hCoV-19/Portugal/PT0568/2020\|EPI_ISL_454291\|2020-0 | ···················· | ···················· | ···················· | ···················· | ···················· | ···················· |
| C | 4239 | hCoV-19/Portugal/PT0569/2020\|EPI_ISL_454292\|2020-0 | ···················· | ···················· | ···················· | ···················· | ···················· | ···················· |
| C | 4240 | hCoV-19/Portugal/PT0570/2020\|EPI_ISL_454293\|2020-0 | ···················· | ···················· | ···················· | ···················· | ···················· | ···················· |
| C | 4241 | hCoV-19/Portugal/PT0571/2020\|EPI_ISL_454294\|2020-0 | ···················· | ···················· | ···················· | ···················· | ···················· | ···················· |
| C | 4242 | hCoV-19/Portugal/PT0572/2020\|EPI_ISL_454295\|2020-0 | ···················· | ···················· | ···················· | ···················· | ···················· | ···················· |
| C | 4243 | hCoV-19/Portugal/PT0573/2020\|EPI_ISL_454296\|2020-0 | ···················· | ···················· | ···················· | ···················· | ···················· | ···················· |
| C | 4244 | hCoV-19/Portugal/PT0574/2020\|EPI_ISL_454297\|2020-0 | ···················· | ···················· | ···················· | ···················· | ···················· | ···················· |
| C | 4245 | hCoV-19/Portugal/PT0575/2020\|EPI_ISL_454298\|2020-0 | ···················· | ···················· | ···················· | ···················· | ···················· | ···················· |
| C | 4246 | hCoV-19/Thailand/Nonthaburi_3481/2020\|EPI_ISL_4555 | ···················· | ···················· | ···················· | ···················· | ···················· | ···················· |
| C | 4247 | hCoV-19/Thailand/Nonthaburi_3480/2020\|EPI_ISL_4555 | ···················· | ···················· | ···················· | ···················· | ···················· | ···················· |
| C | 4248 | hCoV-19/Thailand/Nonthaburi_3479/2020\|EPI_ISL_4555 | ···················· | ···················· | ···················· | ···················· | ···················· | ···················· |
| C | 4249 | hCoV-19/Thailand/Nonthaburi_3478/2020\|EPI_ISL_4555 | ···················· | ···················· | ···················· | ···················· | ···················· | ···················· |
| C | 4250 | hCoV-19/Thailand/Nonthaburi_3472/2020\|EPI_ISL_4555 | ···················· | ···················· | ···················· | ···················· | ···················· | ···················· |
| C | 4251 | hCoV-19/Thailand/Nonthaburi_2983/2020\|EPI_ISL_4555 | ···················· | ···················· | ···················· | ···················· | ···················· | ···················· |
| C | 4252 | hCoV-19/Australia/VPRL002/2020\|EPI_ISL_455595\|2020 | ···················· | ···················· | ···················· | ···················· | ···················· | ···················· |
| C | 4253 | hCoV-19/Australia/VPRL004/2020\|EPI_ISL_455596\|2020 | ···················· | ···················· | ···················· | ···················· | ···················· | ···················· |
| C | 4254 | hCoV-19/Australia/VPRL005/2020\|EPI_ISL_455597\|2020 | ···················· | ···················· | ···················· | ···················· | ···················· | ···················· |
| C | 4255 | hCoV-19/Australia/VPRL006/2020\|EPI_ISL_455598\|2020 | ···················· | ···················· | ···················· | ···················· | ···················· | ···················· |
| C | 4256 | hCoV-19/Australia/VPRL007/2020\|EPI_ISL_455599\|2020 | ···················· | ···················· | ···················· | ···················· | ···················· | ···················· |
| C | 4257 | hCoV-19/Scotland/GCVR-173939/2020\|EPI_ISL_459109\|2 | ···················· | ···················· | ···················· | ···················· | ···················· | ···················· |
| C | 4258 | hCoV-19/Scotland/GCVR-173B51/2020\|EPI_ISL_459111\|2 | ···················· | ···················· | ···················· | ···················· | ···················· | ···················· |
| C | 4259 | hCoV-19/Scotland/GCVR-173CF4/2020\|EPI_ISL_459112\|2 | ···················· | ···················· | ···················· | ···················· | ···················· | ···················· |
| C | 4260 | hCoV-19/Scotland/GCVR-1739EE/2020\|EPI_ISL_459113\|2 | ···················· | ·A·················· | ···················· | ···················· | ···················· | ···················· |
| C | 4261 | hCoV-19/Scotland/GCVR-173D79/2020\|EPI_ISL_459114\|2 | ···················· | ···················· | ···················· | ···················· | ···················· | ···················· |
| C | 4262 | hCoV-19/Scotland/GCVR-173DE2/2020\|EPI_ISL_459115\|2 | ···················· | ···················· | ···················· | ···················· | ···················· | ···················· |
| C | 4263 | hCoV-19/Scotland/GCVR-173D6A/2020\|EPI_ISL_459116\|2 | ···················· | ·A·················· | ···················· | ···················· | ···················· | ···················· |
| C | 4264 | hCoV-19/Scotland/GCVR-173D5B/2020\|EPI_ISL_459118\|2 | ···················· | ···················· | ···················· | ···················· | ···················· | ···················· |
| C | 4265 | hCoV-19/Scotland/GCVR-173D4C/2020\|EPI_ISL_459119\|2 | ···················· | ·A·················· | ···················· | ···················· | ···················· | ···················· |
| C | 4266 | hCoV-19/Scotland/GCVR-173BBB/2020\|EPI_ISL_459120\|2 | ···················· | ···················· | ···················· | ···················· | ···················· | ···················· |
| C | 4267 | hCoV-19/Scotland/GCVR-173C6D/2020\|EPI_ISL_459121\|2 | ···················· | ···················· | ···················· | ···················· | ···················· | ···················· |
| C | 4268 | hCoV-19/Scotland/GCVR-173D3D/2020\|EPI_ISL_459122\|2 | ···················· | ···················· | ···················· | ···················· | ···················· | ···················· |
| C | 4269 | hCoV-19/Scotland/GCVR-173C21/2020\|EPI_ISL_459124\|2 | ···················· | ···················· | ···················· | ···················· | ···················· | ···················· |
| C | 4270 | hCoV-19/Scotland/GCVR-172C04/2020\|EPI_ISL_459125\|2 | ···················· | ·A·················· | ···················· | ···················· | ···················· | ···················· |
| C | 4271 | hCoV-19/Scotland/GCVR-173C4F/2020\|EPI_ISL_459127\|2 | ···················· | ···················· | ···················· | ···················· | ···················· | ···················· |
| C | 4272 | hCoV-19/Scotland/GCVR-173CC7/2020\|EPI_ISL_459130\|2 | ···················· | ···················· | ···················· | ···················· | ···················· | ···················· |
| C | 4273 | hCoV-19/Scotland/GCVR-173E2B/2020\|EPI_ISL_459131\|2 | ···················· | ···················· | ···················· | ···················· | ···················· | ···················· |
| C | 4274 | hCoV-19/Scotland/GCVR-173DD3/2020\|EPI_ISL_459132\|2 | ···················· | ···················· | ···················· | ···················· | ···················· | ···················· |
| C | 4275 | hCoV-19/Scotland/GCVR-173A90/2020\|EPI_ISL_459133\|2 | ···················· | ···················· | ···················· | ···················· | ···················· | ···················· |
| C | 4276 | hCoV-19/Scotland/GCVR-172C22/2020\|EPI_ISL_459134\|2 | ···················· | ···················· | ···················· | ···················· | ···················· | ···················· |
| C | 4277 | hCoV-19/Scotland/GCVR-173A45/2020\|EPI_ISL_459136\|2 | ···················· | ···················· | ···················· | ···················· | ···················· | ···················· |
| C | 4278 | hCoV-19/Scotland/GCVR-172B9E/2020\|EPI_ISL_459137\|2 | ···················· | ···················· | ···················· | ···················· | ···················· | ···················· |
| C | 4279 | hCoV-19/Scotland/GCVR-1739FD/2020\|EPI_ISL_459138\|2 | ···················· | ··············C····· | ···················· | ···················· | ···················· | ···················· |
| C | 4280 | hCoV-19/Scotland/GCVR-173D97/2020\|EPI_ISL_459140\|2 | ···················· | ···················· | ···················· | ···················· | ···················· | ···················· |
| C | 4281 | hCoV-19/Scotland/GCVR-172BF8/2020\|EPI_ISL_459141\|2 | ···················· | ···················· | ···················· | ···················· | ···················· | ···················· |
| C | 4282 | hCoV-19/Scotland/GCVR-173BAC/2020\|EPI_ISL_459142\|2 | ···················· | ···················· | ···················· | ···················· | ···················· | ···················· |
| C | 4283 | hCoV-19/Scotland/GCVR-173B60/2020\|EPI_ISL_459145\|2 | ···················· | ···················· | ···················· | ···················· | ···················· | ···················· |
| C | 4284 | hCoV-19/Scotland/GCVR-173BF7/2020\|EPI_ISL_459146\|2 | ···················· | ···················· | ···················· | ···················· | ···················· | ···················· |
| C | 4285 | hCoV-19/Scotland/GCVR-173CD6/2020\|EPI_ISL_459147\|2 | ···················· | ···················· | ···················· | ···················· | ···················· | ···················· |
| C | 4286 | hCoV-19/Scotland/GCVR-173A81/2020\|EPI_ISL_459149\|2 | ···················· | ···················· | ···················· | ···················· | ···················· | ···················· |
| C | 4287 | hCoV-19/USA/MA-MGH-00016/2020\|EPI_ISL_460101\|2020- | ···················· | ···················· | ···················· | ···················· | ···················· | ···················· |
| C | 4288 | hCoV-19/USA/MA-MGH-00018/2020\|EPI_ISL_460102\|2020- | ···················· | ···················· | ···················· | ···················· | ···················· | ···················· |
| C | 4289 | hCoV-19/USA/MA-MGH-00426/2020\|EPI_ISL_460103\|2020- | ···················· | ···················· | ···················· | ···················· | ···················· | ···················· |
| C | 4290 | hCoV-19/USA/MA-MGH-00050/2020\|EPI_ISL_460104\|2020- | ···················· | ···················· | ···················· | ···················· | ···················· | ···················· |
| C | 4291 | hCoV-19/USA/MA-MGH-00042/2020\|EPI_ISL_460105\|2020- | ···················· | ···················· | ···················· | ···················· | ···················· | ···················· |
| C | 4292 | hCoV-19/USA/MA-MGH-00707/2020\|EPI_ISL_460106\|2020- | ···················· | ···················· | ···················· | ···················· | ···················· | ···················· |
| C | 4293 | hCoV-19/USA/MA-MGH-00157/2020\|EPI_ISL_460107\|2020- | ···················· | ···················· | ···················· | ···················· | ···················· | ···················· |
| C | 4294 | hCoV-19/USA/MA-MGH-00208/2020\|EPI_ISL_460108\|2020- | ···················· | ···················· | ···················· | ···················· | ···················· | ···················· |
| C | 4295 | hCoV-19/USA/MA-MGH-00067/2020\|EPI_ISL_460109\|2020- | ···················· | ···················· | ···················· | ···················· | ···················· | ···················· |
| C | 4296 | hCoV-19/USA/MA-MGH-00485/2020\|EPI_ISL_460110\|2020- | ···················· | ···················· | ···················· | ···················· | ···················· | ···················· |
| C | 4297 | hCoV-19/USA/MA-MGH-00031/2020\|EPI_ISL_460111\|2020- | ···················· | ···················· | ···················· | ···················· | ···················· | ···················· |
| C | 4298 | hCoV-19/USA/MA-MGH-00708/2020\|EPI_ISL_460112\|2020- | ···················· | ···················· | ···················· | ···················· | ···················· | ···················· |
| C | 4299 | hCoV-19/USA/MA-MGH-00143/2020\|EPI_ISL_460114\|2020- | ···················· | ···················· | ···················· | ···················· | ···················· | ···················· |
| C | 4300 | hCoV-19/USA/MA-MGH-00588/2020\|EPI_ISL_460115\|2020- | ···················· | ···················· | ···················· | ···················· | ···················· | ···················· |
| C | 4301 | hCoV-19/USA/MA-MGH-00440/2020\|EPI_ISL_460116\|2020- | ···················· | ···················· | ···················· | ···················· | ···················· | ···················· |
| C | 4302 | hCoV-19/USA/MA-MGH-00417/2020\|EPI_ISL_460117\|2020- | ···················· | ···················· | ···················· | ···················· | ···················· | ···················· |
| C | 4303 | hCoV-19/USA/MA-MGH-00678/2020\|EPI_ISL_460118\|2020- | ···················· | ···················· | ···················· | ···················· | ···················· | ···················· |
| C | 4304 | hCoV-19/USA/MA-MGH-00069/2020\|EPI_ISL_460119\|2020- | ···················· | ···················· | ···················· | ···················· | ···················· | ···················· |
| C | 4305 | hCoV-19/USA/MA-MGH-00656/2020\|EPI_ISL_460120\|2020- | ···················· | ···················· | ···················· | ···················· | ···················· | ···················· |
| C | 4306 | hCoV-19/USA/MA-MGH-00258/2020\|EPI_ISL_460121\|2020- | ···················· | ···················· | ···················· | ···················· | ···················· | ···················· |
| C | 4307 | hCoV-19/USA/MA-MGH-00080/2020\|EPI_ISL_460122\|2020- | ···················· | ···················· | ···················· | ···················· | ···················· | ···················· |
| C | 4308 | hCoV-19/USA/MA-MGH-00711/2020\|EPI_ISL_460123\|2020- | ···················· | ···················· | ···················· | ···················· | ···················· | ···················· |
| C | 4309 | hCoV-19/USA/MA-MGH-00182/2020\|EPI_ISL_460124\|2020- | ···················· | ···················· | ···················· | ···················· | ···················· | ···················· |
| C | 4310 | hCoV-19/USA/MA-MGH-00562/2020\|EPI_ISL_460125\|2020- | ···················· | ···················· | ···················· | ···················· | ···················· | ···················· |
| C | 4311 | hCoV-19/USA/MA-MGH-00065/2020\|EPI_ISL_460126\|2020- | ···················· | ···················· | ···················· | ···················· | ···················· | ···················· |
| C | 4312 | hCoV-19/USA/MA-MGH-00100/2020\|EPI_ISL_460128\|2020- | ···················· | ···················· | ···················· | ···················· | ···················· | ···················· |
| C | 4313 | hCoV-19/USA/MA-MGH-00627/2020\|EPI_ISL_460129\|2020- | ···················· | ···················· | ···················· | ···················· | ···················· | ···················· |

| | | | | | | | | | |
|---|---|---|---|---|---|---|---|---|---|
| C | 4314 | hCoV-19/USA/MA-MGH-00706/2020\|EPI_ISL_460130\|2020- | .................... | .................... | .................... | .................... | .................... | .................... | |
| C | 4315 | hCoV-19/USA/MA-MGH-00267/2020\|EPI_ISL_460131\|2020- | .................... | .................... | .................... | .................... | .................... | .................... | |
| C | 4316 | hCoV-19/USA/CT-UW-6603/2020\|EPI_ISL_461420\|2020-04 | .................... | .................... | .................... | .................... | .................... | .................... | |
| C | 4317 | hCoV-19/USA/WA-UW-6605/2020\|EPI_ISL_461421\|2020-04 | .................... | .................... | .................... | .................... | .................... | .................... | |
| C | 4318 | hCoV-19/USA/WA-UW-3904/2020\|EPI_ISL_461422\|2020-03 | .................... | .................... | .................... | .................... | .................... | .................... | |
| C | 4319 | hCoV-19/USA/WA-UW-4117/2020\|EPI_ISL_461423\|2020-03 | .................... | .................... | .................... | .................... | .................... | .................... | |
| C | 4320 | hCoV-19/USA/ID-UW-4376/2020\|EPI_ISL_461424\|2020-03 | .................... | .................... | .................... | .................... | .................... | .................... | |
| C | 4321 | hCoV-19/USA/ID-UW-4595/2020\|EPI_ISL_461425\|2020-03 | .................... | .................... | .................... | .................... | .................... | .................... | |
| C | 4322 | hCoV-19/USA/WA-UW-4659/2020\|EPI_ISL_461426\|2020-04 | .................... | .................... | .................... | .................... | .................... | .................... | |
| C | 4323 | hCoV-19/USA/WA-UW-4676/2020\|EPI_ISL_461427\|2020-04 | .................... | .................... | .................... | .................... | .................... | .................... | |
| C | 4324 | hCoV-19/USA/WA-UW-4688/2020\|EPI_ISL_461428\|2020-04 | .................... | .................... | .................... | .................... | .................... | .................... | |
| C | 4325 | hCoV-19/USA/WA-UW-4690/2020\|EPI_ISL_461429\|2020-04 | .................... | .................... | .................... | .................... | .................... | .................... | |
| C | 4326 | hCoV-19/USA/WA-UW-4746/2020\|EPI_ISL_461430\|2020-03 | .................... | .................... | .................... | .................... | .................... | .................... | |
| C | 4327 | hCoV-19/USA/WA-UW-4779/2020\|EPI_ISL_461432\|2020-03 | .................... | .................... | .................... | .................... | .................... | .................... | |
| C | 4328 | hCoV-19/USA/WA-UW-5016/2020\|EPI_ISL_461434\|2020-04 | .................... | .................... | .................... | .................... | .................... | .................... | |
| C | 4329 | hCoV-19/USA/CT-UW-5104/2020\|EPI_ISL_461435\|2020-04 | .................... | .................... | .................... | .................... | .................... | .................... | |
| C | 4330 | hCoV-19/USA/WA-UW-5455/2020\|EPI_ISL_461436\|2020-03 | .................... | .................... | .................... | .................... | .................... | .................... | |
| C | 4331 | hCoV-19/USA/UN-UW-5620/2020\|EPI_ISL_461437\|2020-04 | .................... | .................... | .................... | .................... | .................... | .................... | |
| C | 4332 | hCoV-19/USA/UN-UW-5622/2020\|EPI_ISL_461438\|2020-04 | .................... | .................... | .................... | .................... | .................... | .................... | |
| C | 4333 | hCoV-19/USA/WA-UW-5738/2020\|EPI_ISL_461440\|2020-04 | .................... | .................... | .................... | .................... | .................... | .................... | |
| C | 4334 | hCoV-19/USA/CT-UW-5765/2020\|EPI_ISL_461441\|2020-04 | .................... | .................... | .................... | .................... | .................... | .................... | |
| C | 4335 | hCoV-19/USA/WA-UW-5781/2020\|EPI_ISL_461442\|2020-04 | .................... | .................... | .................... | .................... | .................... | .................... | |
| C | 4336 | hCoV-19/USA/WA-UW-5847/2020\|EPI_ISL_461444\|2020-03 | .................... | .................... | .................... | .................... | .................... | .................... | |
| C | 4337 | hCoV-19/USA/WA-UW-5860/2020\|EPI_ISL_461445\|2020-03 | .................... | .................... | .................... | .................... | .................... | .................... | |
| C | 4338 | hCoV-19/USA/WA-UW-6663/2020\|EPI_ISL_461446\|2020-04 | .................... | .................... | .................... | .................... | .................... | .................... | |
| C | 4339 | hCoV-19/USA/WA-UW-6671/2020\|EPI_ISL_461447\|2020-04 | .................... | .................... | .................... | .................... | .................... | .................... | |
| C | 4340 | hCoV-19/USA/WA-UW-6698/2020\|EPI_ISL_461448\|2020-04 | .................... | .................... | .................... | .................... | .................... | .................... | |
| C | 4341 | hCoV-19/USA/WA-UW-6701/2020\|EPI_ISL_461449\|2020-04 | .................... | .................... | .................... | .................... | .................... | .................... | |
| C | 4342 | hCoV-19/USA/WA-UW-10027/2020\|EPI_ISL_461450\|2020-0 | .................... | .................... | .................... | .................... | .................... | .................... | |
| C | 4343 | hCoV-19/USA/WA-UW-10028/2020\|EPI_ISL_461451\|2020-0 | .................... | .................... | .................... | .................... | .................... | .................... | |
| C | 4344 | hCoV-19/USA/WA-UW-10029/2020\|EPI_ISL_461452\|2020-0 | .................... | .................... | .................... | .................... | .................... | .................... | |
| C | 4345 | hCoV-19/USA/WA-UW-10031/2020\|EPI_ISL_461453\|2020-0 | .................... | .................... | .................... | .................... | .................... | .................... | |
| C | 4346 | hCoV-19/USA/WA-UW-10036/2020\|EPI_ISL_461454\|2020-0 | .................... | .................... | .................... | .................... | .................... | .................... | |
| C | 4347 | hCoV-19/USA/WA-UW-10039/2020\|EPI_ISL_461455\|2020-0 | .................... | .................... | .................... | .................... | .................... | .................... | |
| C | 4348 | hCoV-19/USA/WA-UW-10040/2020\|EPI_ISL_461456\|2020-0 | .................... | .................... | .................... | .................... | .................... | .................... | |
| C | 4349 | hCoV-19/USA/WA-UW-10042/2020\|EPI_ISL_461457\|2020-0 | .................... | .................... | .................... | .................... | .................... | .................... | |
| C | 4350 | hCoV-19/USA/WA-UW-10088/2020\|EPI_ISL_461458\|2020-0 | .................... | .................... | .................... | .................... | .................... | .................... | |
| C | 4351 | hCoV-19/USA/WA-UW-10089/2020\|EPI_ISL_461459\|2020-0 | .................... | .................... | .................... | .................... | .................... | .................... | |
| C | 4352 | hCoV-19/USA/WA-UW-10090/2020\|EPI_ISL_461460\|2020-0 | .................... | .................... | .................... | .................... | .................... | .................... | |
| C | 4353 | hCoV-19/Israel/701002555/2020\|EPI_ISL_447290\|2020- | .................... | .................... | .................... | .................... | .................... | .................... | |
| C | 4354 | hCoV-19/Israel/701002550/2020\|EPI_ISL_447291\|2020- | .................... | .................... | .................... | .................... | .................... | .................... | |
| C | 4355 | hCoV-19/Israel/701002540/2020\|EPI_ISL_447292\|2020- | .................... | .................... | .................... | .................... | .................... | .................... | |
| C | 4356 | hCoV-19/Israel/701002538/2020\|EPI_ISL_447293\|2020- | .................... | .................... | .................... | .................... | .................... | .................... | |
| C | 4357 | hCoV-19/Israel/701002504/2020\|EPI_ISL_447294\|2020- | .................... | .................... | .................... | .................... | .................... | .................... | |
| C | 4358 | hCoV-19/Israel/701002489/2020\|EPI_ISL_447295\|2020- | .................... | .................... | .................... | .................... | .................... | .................... | |
| C | 4359 | hCoV-19/Israel/701002462/2020\|EPI_ISL_447296\|2020- | .................... | .................... | .................... | .................... | .................... | .................... | |
| C | 4360 | hCoV-19/Israel/701002458/2020\|EPI_ISL_447297\|2020- | .................... | .................... | .................... | .................... | .................... | .................... | |
| C | 4361 | hCoV-19/Israel/701002455/2020\|EPI_ISL_447299\|2020- | .................... | .................... | .................... | .................... | .................... | .................... | |
| C | 4362 | hCoV-19/England/LOND-D55F6/2020\|EPI_ISL_449704\|202 | .................... | .................... | .................... | .................... | .................... | .................... | |
| C | 4363 | hCoV-19/England/LOND-D5602/2020\|EPI_ISL_449705\|202 | .................... | .................... | .................... | .................... | .................... | .................... | |
| C | 4364 | hCoV-19/England/LOND-D5611/2020\|EPI_ISL_449706\|202 | .................... | .................... | .................... | .................... | .................... | .................... | |
| C | 4365 | hCoV-19/England/LOND-D5620/2020\|EPI_ISL_449707\|202 | .................... | .................... | .................... | .................... | .................... | .................... | |
| C | 4366 | hCoV-19/England/LOND-D564E/2020\|EPI_ISL_449708\|202 | .................... | .................... | .................... | .................... | .................... | .................... | |
| C | 4367 | hCoV-19/England/LOND-D565D/2020\|EPI_ISL_449709\|202 | .................... | .................... | .................... | .................... | .................... | .................... | |
| C | 4368 | hCoV-19/England/LOND-D56E4/2020\|EPI_ISL_449715\|202 | .................... | .................... | .................... | .................... | .................... | .................... | |
| C | 4369 | hCoV-19/England/LOND-D56F3/2020\|EPI_ISL_449716\|202 | .................... | .................... | .................... | .................... | .................... | .................... | |
| C | 4370 | hCoV-19/England/LOND-D574B/2020\|EPI_ISL_449718\|202 | .................... | .................... | .................... | .................... | .................... | .................... | |
| C | 4371 | hCoV-19/England/LOND-D5778/2020\|EPI_ISL_449719\|202 | .................... | .................... | .................... | .................... | .................... | .................... | |
| C | 4372 | hCoV-19/England/LOND-D5796/2020\|EPI_ISL_449720\|202 | .................... | .................... | .................... | .................... | .................... | .................... | |
| C | 4373 | hCoV-19/England/LOND-D57A5/2020\|EPI_ISL_449721\|202 | .................... | .................... | .................... | .................... | .................... | .................... | |
| C | 4374 | hCoV-19/England/LOND-D57B4/2020\|EPI_ISL_449722\|202 | .................... | .................... | .................... | .................... | .................... | .................... | |
| C | 4375 | hCoV-19/England/LOND-D57C3/2020\|EPI_ISL_449723\|202 | .................... | .................... | .................... | .................... | .................... | .................... | |
| C | 4376 | hCoV-19/England/LOND-D57E1/2020\|EPI_ISL_449724\|202 | .................... | .................... | .................... | .................... | .................... | .................... | |
| C | 4377 | hCoV-19/England/LOND-D57F0/2020\|EPI_ISL_449725\|202 | .................... | .................... | .................... | .................... | .................... | .................... | |
| C | 4378 | hCoV-19/USA/WA-S730/2020\|EPI_ISL_449900\|2020-04-11 | .................... | .................... | .................... | .................... | .................... | .................... | |
| C | 4379 | hCoV-19/USA/WA-S731/2020\|EPI_ISL_449901\|2020-04-11 | .................... | .................... | .................... | .................... | .................... | .................... | |
| C | 4380 | hCoV-19/USA/WI-UW-348/2020\|EPI_ISL_450702\|2020-04- | .................... | .................... | .................... | .................... | .................... | .................... | |
| C | 4381 | hCoV-19/USA/IL-UW-349/2020\|EPI_ISL_450703\|2020-04- | .................... | .................... | .................... | .................... | .................... | .................... | |
| C | 4382 | hCoV-19/USA/IL-UW-350/2020\|EPI_ISL_450704\|2020-04- | .................... | .................... | .................... | .................... | .................... | .................... | |
| C | 4383 | hCoV-19/USA/WI-UW-351/2020\|EPI_ISL_450705\|2020-04- | .................... | .................... | .................... | .................... | .................... | .................... | |
| C | 4384 | hCoV-19/USA/IL-UW-352/2020\|EPI_ISL_450706\|2020-04- | .................... | .................... | .................... | .................... | .................... | .................... | |
| C | 4385 | hCoV-19/USA/IL-UW-353/2020\|EPI_ISL_450707\|2020-04- | .................... | .................... | .................... | .................... | .................... | .................... | |
| C | 4386 | hCoV-19/USA/WI-UW-354/2020\|EPI_ISL_450708\|2020-04- | .................... | .................... | .................... | .................... | .................... | .................... | |
| C | 4387 | hCoV-19/USA/IL-UW-355/2020\|EPI_ISL_450709\|2020-04- | .................... | .................... | .................... | .................... | .................... | .................... | |
| C | 4388 | hCoV-19/Scotland/EDB3662/2020\|EPI_ISL_453101\|2020- | .................... | .A.................. | .................... | .................... | .................... | .................... | |
| C | 4389 | hCoV-19/Scotland/EDB3991/2020\|EPI_ISL_453102\|2020- | .................... | ...................A | .................... | .................... | .................... | .................... | 1 |
| C | 4390 | hCoV-19/Scotland/EDB3992/2020\|EPI_ISL_453103\|2020- | .................... | ...................A | .................... | .................... | .................... | .................... | 1 |
| C | 4391 | hCoV-19/Scotland/EDB4006/2020\|EPI_ISL_453104\|2020- | .................... | .................... | .................... | .................... | .................... | .................... | |

| | | | | | | | | | |
|---|---|---|---|---|---|---|---|---|---|
| C | 4392 | hCoV-19/Scotland/EDB4012/2020\|EPI_ISL_453105\|2020- | ···················· | ···················· | ···················· | ···················· | ···················· | ···················· | |
| C | 4393 | hCoV-19/Scotland/EDB4024/2020\|EPI_ISL_453106\|2020- | ···················· | ···················· | ···················· | ···················· | ···················· | ···················· | |
| C | 4394 | hCoV-19/Scotland/EDB4027/2020\|EPI_ISL_453107\|2020- | ···················· | ···················A | ···················· | ···················· | ···················· | ···················· | 1 |
| C | 4395 | hCoV-19/Scotland/EDB4113/2020\|EPI_ISL_453108\|2020- | ···················· | ···················A | ···················· | ···················· | ···················· | ···················· | 1 |
| C | 4396 | hCoV-19/Scotland/EDB4176/2020\|EPI_ISL_453109\|2020- | ···················· | ···················· | ···················· | ···················· | ···················· | ···················· | |
| C | 4397 | hCoV-19/Scotland/EDB4181/2020\|EPI_ISL_453111\|2020- | ···················· | ·A·················· | ···················· | ···················· | ···················· | ···················· | |
| C | 4398 | hCoV-19/Scotland/EDB4233/2020\|EPI_ISL_453114\|2020- | ···················· | ···················A | ···················· | ···················· | ··············N····· | ···················· | |
| C | 4399 | hCoV-19/Scotland/EDB4234/2020\|EPI_ISL_453115\|2020- | ···················· | ···················A | ···················· | ···················· | ···················· | ···················· | 1 |
| C | 4400 | hCoV-19/Scotland/EDB4235/2020\|EPI_ISL_453116\|2020- | ···················· | ···················A | ···················· | ···················· | ···················· | ···················· | 1 |
| C | 4402 | hCoV-19/Scotland/EDB4305/2020\|EPI_ISL_453119\|2020- | ···················· | ···················A | ···················· | ···················· | ···················· | ···················· | 1 |
| C | 4403 | hCoV-19/Scotland/EDB4329/2020\|EPI_ISL_453121\|2020- | ···················· | ···················A | ···················· | ···················· | ···················· | ···················· | 1 |
| C | 4404 | hCoV-19/Scotland/EDB4335/2020\|EPI_ISL_453122\|2020- | ···················· | ···················· | ···················· | ···················· | ···················· | ···················· | |
| C | 4405 | hCoV-19/Scotland/EDB4357/2020\|EPI_ISL_453125\|2020- | ···················· | ·A·················· | ···················· | ···················· | ···················· | ···················· | |
| C | 4406 | hCoV-19/Scotland/EDB4360/2020\|EPI_ISL_453126\|2020- | ···················· | ···················A | ···················· | ···················· | ···················· | ···················· | 1 |
| C | 4407 | hCoV-19/Scotland/EDB4363/2020\|EPI_ISL_453127\|2020- | ···················· | ···················· | ···················· | ···················· | ···················· | ···················· | |
| C | 4408 | hCoV-19/Scotland/EDB4364/2020\|EPI_ISL_453128\|2020- | ···················· | ···················· | ···················· | ···················· | ···················· | ···················· | |
| C | 4409 | hCoV-19/Scotland/EDB4453/2020\|EPI_ISL_453129\|2020- | ···················· | ·A·················· | ···················· | ···················· | ···················· | ···················· | |
| C | 4410 | hCoV-19/Scotland/EDB4628/2020\|EPI_ISL_453130\|2020- | ···················· | ·A·················· | ···················· | ···················· | ···················· | ···················· | |
| C | 4411 | hCoV-19/Scotland/EDB4676/2020\|EPI_ISL_453132\|2020- | ···················· | ·A·················· | ···················· | ···················· | ···················· | ···················· | |
| C | 4412 | hCoV-19/Scotland/EDB4716/2020\|EPI_ISL_453133\|2020- | ···················· | ···················· | ···················· | ···················· | ···················· | ···················· | |
| C | 4413 | hCoV-19/Scotland/EDB4738/2020\|EPI_ISL_453141\|2020- | ···················· | ···················· | ···················· | ···················· | ···················· | ···················· | |
| C | 4414 | hCoV-19/Scotland/EDB4739/2020\|EPI_ISL_453142\|2020- | ···················· | ···················· | ···················· | ···················· | ···················· | ···················· | |
| C | 4415 | hCoV-19/Scotland/EDB4743/2020\|EPI_ISL_453143\|2020- | ···················· | ···················· | ···················· | ···················· | ···················· | ···················· | |
| C | 4416 | hCoV-19/Scotland/EDB4744/2020\|EPI_ISL_453144\|2020- | ···················· | ·A·················· | ···················· | ···················· | ···················· | ···················· | |
| C | 4417 | hCoV-19/Scotland/EDB4745/2020\|EPI_ISL_453145\|2020- | ···················· | ·A·················· | ···················· | ···················· | ···················· | ···················· | |
| C | 4418 | hCoV-19/Scotland/EDB4746/2020\|EPI_ISL_453146\|2020- | ···················· | ·A·················· | ···················· | ···················· | ···················· | ···················· | |
| C | 4419 | hCoV-19/Scotland/EDB4752/2020\|EPI_ISL_453147\|2020- | ···················· | ···················· | ···················· | ···················· | ···················· | ···················· | |
| C | 4421 | hCoV-19/Scotland/EDB4756/2020\|EPI_ISL_453149\|2020- | ···················· | ·A·················· | ···················· | ···················· | ···················· | ···················· | |
| C | 4422 | hCoV-19/Scotland/EDB4757/2020\|EPI_ISL_453150\|2020- | ···················· | ·A·················· | ···················· | ···················· | ···················· | ···················· | |
| C | 4423 | hCoV-19/Scotland/EDB4800/2020\|EPI_ISL_453152\|2020- | ···················· | ···················· | ···················· | ···················· | ···················· | ···················· | |
| C | 4424 | hCoV-19/Scotland/EDB4802/2020\|EPI_ISL_453153\|2020- | ···················· | ···················· | ···················· | ···················· | ···················· | ···················· | |
| C | 4425 | hCoV-19/Scotland/EDB4806/2020\|EPI_ISL_453154\|2020- | ···················· | ·A·················· | ···················· | ···················· | ···················· | ···················· | |
| C | 4426 | hCoV-19/Scotland/EDB4822/2020\|EPI_ISL_453155\|2020- | ···················· | ·A·················· | ···················· | ···················· | ···················· | ···················· | |
| C | 4427 | hCoV-19/Scotland/EDB4823/2020\|EPI_ISL_453156\|2020- | ···················· | ···················A | ···················· | ···················· | ···················· | ···················· | 1 |
| C | 4428 | hCoV-19/Scotland/EDB4828/2020\|EPI_ISL_453157\|2020- | ···················· | ···················· | ···················· | ···················· | ···················· | ···················· | |
| C | 4429 | hCoV-19/Scotland/EDB4830/2020\|EPI_ISL_453158\|2020- | ···················· | ···················· | ···················· | ···················· | ···················· | ···················· | |
| C | 4431 | hCoV-19/Scotland/EDB4835/2020\|EPI_ISL_453161\|2020- | ···················· | ···················· | ···················· | ···················· | ···················· | ···················· | |
| C | 4432 | hCoV-19/Scotland/EDB4838/2020\|EPI_ISL_453163\|2020- | ···················· | ···················A | ···················· | ···················· | ···················· | ···················· | 1 |
| C | 4433 | hCoV-19/Scotland/EDB4839/2020\|EPI_ISL_453164\|2020- | ···················· | ···················A | ···················· | ···················· | ···················· | ···················· | 1 |
| C | 4434 | hCoV-19/Scotland/EDB4840/2020\|EPI_ISL_453165\|2020- | ···················· | ···················· | ···················· | ···················· | ···················· | ···················· | |
| C | 4435 | hCoV-19/Scotland/EDB4842/2020\|EPI_ISL_453166\|2020- | ···················· | ···················· | ···················· | ···················· | ···················· | ···················· | |
| C | 4436 | hCoV-19/USA/PA-MGEL137/2020\|EPI_ISL_454401\|2020-04 | ···················· | ···················· | ···················· | ···················· | ···················· | ···················· | |
| C | 4437 | hCoV-19/USA/PA-MGEL140/2020\|EPI_ISL_454402\|2020-04 | ···················· | ···················· | ···················· | ···················· | ···················· | ···················· | |
| C | 4438 | hCoV-19/USA/PA-MGEL143/2020\|EPI_ISL_454403\|2020-04 | ···················· | ···················· | ···················· | ···················· | ···················· | ···················· | |
| C | 4439 | hCoV-19/USA/PA-MGEL146/2020\|EPI_ISL_454404\|2020-04 | ···················· | ···················· | ···················· | ···················· | ···················· | ···················· | |
| C | 4440 | hCoV-19/USA/PA-MGEL147/2020\|EPI_ISL_454405\|2020 | ···················· | ···················· | ···················· | ···················· | ···················· | ···················· | |
| C | 4441 | hCoV-19/USA/PA-MGEL148/2020\|EPI_ISL_454406\|2020-04 | ···················· | ···················· | ···················· | ···················· | ···················· | ···················· | |
| C | 4442 | hCoV-19/USA/PA-MGEL151/2020\|EPI_ISL_454408\|2020-04 | ···················· | ···················· | ···················· | ···················· | ···················· | ···················· | |
| C | 4443 | hCoV-19/USA/PA-MGEL152/2020\|EPI_ISL_454409\|2020-04 | ···················· | ···················· | ···················· | ···················· | ···················· | ···················· | |
| C | 4444 | hCoV-19/USA/NC_200213/2020\|EPI_ISL_454412\|2020-03 | ···················· | ···················· | ···················· | ···················· | ···················· | ···················· | |
| C | 4445 | hCoV-19/USA/NC_200214/2020\|EPI_ISL_454413\|2020-03 | ···················· | ···················· | ···················· | ···················· | ···················· | ···················· | |
| C | 4446 | hCoV-19/USA/NC_200215/2020\|EPI_ISL_454414\|2020-03 | ···················· | ···················· | ···················· | ···················· | ···················· | ···················· | |
| C | 4447 | hCoV-19/USA/NC_200216/2020\|EPI_ISL_454415\|2020-03 | ···················· | ···················· | ···················· | ···················· | ···················· | ···················· | |
| C | 4448 | hCoV-19/Netherlands/Leiden-0002/2020\|EPI_ISL_45441 | ···················· | ···················· | ···················· | ···················· | ···················· | ···················· | |
| C | 4449 | hCoV-19/Guangdong/MZ01/2020\|EPI_ISL_454417\|2020-02 | ···················· | ···················· | ···················· | ···················· | ···················· | ···················· | |
| C | 4450 | hCoV-19/Guangdong/MZ02/2020\|EPI_ISL_454418\|2020-02 | ···················· | ···················· | ···················· | ···················· | ···················· | ···················· | |
| C | 4451 | hCoV-19/USA/MDH-8144/2020\|EPI_ISL_454431\|2020-03-1 | ···················· | ···················· | ···················· | ···················· | ···················· | ···················· | |
| C | 4452 | hCoV-19/USA/MDH-9483/2020\|EPI_ISL_454435\|2020-03-1 | ···················· | ···················· | ···················· | ···················· | ···················· | ···················· | |
| C | 4453 | hCoV-19/Sweden/20-08040/2020\|EPI_ISL_454436\|2020-0 | ···················· | ···················· | ···················· | ···················· | ···················· | ···················· | |
| C | 4454 | hCoV-19/Sweden/20-08042/2020\|EPI_ISL_454437\|2020-0 | ···················· | ···················· | ···················· | ···················· | ···················· | ···················· | |
| C | 4455 | hCoV-19/Sweden/20-08043/2020\|EPI_ISL_454438\|2020-0 | ···················· | ···················· | ···················· | ···················· | ···················· | ···················· | |
| C | 4456 | hCoV-19/Sweden/20-08044/2020\|EPI_ISL_454439\|2020-0 | ···················· | ···················· | ···················· | ···················· | ···················· | ···················· | |
| C | 4457 | hCoV-19/Sweden/20-08124/2020\|EPI_ISL_454440\|2020-0 | ···················· | ···················· | ···················· | ···················· | ···················· | ···················· | |
| C | 4458 | hCoV-19/Sweden/20-08125/2020\|EPI_ISL_454441\|2020-0 | ···················· | ···················· | ···················· | ···················· | ···················· | ···················· | |
| C | 4459 | hCoV-19/Sweden/20-08126/2020\|EPI_ISL_454442\|2020-0 | ···················· | ···················· | ···················· | ···················· | ···················· | ···················· | |
| C | 4460 | hCoV-19/Sweden/20-08128/2020\|EPI_ISL_454443\|2020-0 | ···················· | ···················· | ···················· | ···················· | ···················· | ···················· | |
| C | 4461 | hCoV-19/Sweden/20-08129/2020\|EPI_ISL_454444\|2020-0 | ···················· | ···················· | ···················· | ···················· | ···················· | ···················· | |
| C | 4462 | hCoV-19/Sweden/20-08131/2020\|EPI_ISL_454445\|2020-0 | ···················· | ···················· | ···················· | ···················· | ···················· | ···················· | |
| C | 4463 | hCoV-19/Sweden/20-08132/2020\|EPI_ISL_454446\|2020-0 | ···················· | ···················· | ···················· | ···················· | ···················· | ···················· | |
| C | 4464 | hCoV-19/Sweden/20-08060/2020\|EPI_ISL_454447\|2020-0 | ···················· | ···················· | ···················· | ···················· | ···················· | ···················· | |
| C | 4465 | hCoV-19/Sweden/20-08071/2020\|EPI_ISL_454448\|2020-0 | ···················· | ···················· | ···················· | ···················· | ···················· | ···················· | |
| C | 4466 | hCoV-19/Sweden/20-08072/2020\|EPI_ISL_454449\|2020-0 | ···················· | ···················· | ···················· | ···················· | ···················· | ···················· | |
| C | 4467 | hCoV-19/Sweden/20-08073/2020\|EPI_ISL_454450\|2020-0 | ···················· | ···················· | ···················· | ···················· | ···················· | ···················· | |
| C | 4468 | hCoV-19/Sweden/20-08076/2020\|EPI_ISL_454451\|2020-0 | ···················· | ···················· | ···················· | ···················· | ···················· | ···················· | |
| C | 4469 | hCoV-19/Sweden/20-08077/2020\|EPI_ISL_454452\|2020-0 | ···················· | ···················· | ···················· | ···················· | ···················· | ···················· | |
| C | 4470 | hCoV-19/Sweden/20-08078/2020\|EPI_ISL_454453\|2020-0 | ···················· | ···················· | ···················· | ···················· | ···················· | ···················· | |
| C | 4471 | hCoV-19/Sweden/20-08079/2020\|EPI_ISL_454454\|2020-0 | ···················· | ···················· | ···················· | ···················· | ···················· | ···················· | |
| C | 4472 | hCoV-19/Sweden/20-08081/2020\|EPI_ISL_454455\|2020-0 | ···················· | ···················· | ···················· | ···················· | ···················· | ···················· | |

C 4473 hCoV-19/Sweden/20-08082/2020|EPI_ISL_454456|2020-0 ···················· ···················· ···················· ···················· ···················· ····················
C 4474 hCoV-19/Sweden/20-08083/2020|EPI_ISL_454457|2020-0 ···················· ···················· ···················· ···················· ···················· ····················
C 4475 hCoV-19/Sweden/20-08084/2020|EPI_ISL_454458|2020-0 ···················· ···················· ···················· ···················· ···················· ····················
C 4476 hCoV-19/Sweden/20-08085/2020|EPI_ISL_454459|2020-0 ···················· ···················· ···················· ···················· ···················· ····················
C 4477 hCoV-19/Sweden/20-08086/2020|EPI_ISL_454460|2020-0 ···················· ···················· ···················· ···················· ···················· ····················
C 4478 hCoV-19/Sweden/20-08088/2020|EPI_ISL_454461|2020-0 ···················· ···················· ···················· ···················· ···················· ····················
C 4479 hCoV-19/Sweden/20-08090/2020|EPI_ISL_454462|2020-0 ···················· ···················· ···················· ···················· ···················· ····················
C 4480 hCoV-19/Sweden/20-08091/2020|EPI_ISL_454463|2020-0 ···················· ···················· ···················· ···················· ···················· ····················
C 4481 hCoV-19/Sweden/20-08092/2020|EPI_ISL_454464|2020-0 ···················· ···················· ···················· ···················· ···················· ····················
C 4482 hCoV-19/Sweden/20-08093/2020|EPI_ISL_454465|2020-0 ···················· ···················· ···················· ···················· ···················· ····················
C 4483 hCoV-19/Sweden/20-08094/2020|EPI_ISL_454466|2020-0 ···················· ···················· ···················· ···················· ···················· ····················
C 4484 hCoV-19/Sweden/20-08095/2020|EPI_ISL_454467|2020-0 ···················· ···················· ···················· ···················· ···················· ····················
C 4485 hCoV-19/Sweden/20-08096/2020|EPI_ISL_454468|2020-0 ···················· ···················· ···················· ···················· ···················· ····················
C 4486 hCoV-19/Sweden/20-08098/2020|EPI_ISL_454469|2020-0 ···················· ···················· ···················· ···················· ···················· ····················
C 4487 hCoV-19/Sweden/20-08099/2020|EPI_ISL_454470|2020-0 ···················· ···················· ···················· ···················· ···················· ····················
C 4488 hCoV-19/Sweden/20-08100/2020|EPI_ISL_454471|2020-0 ···················· ···················· ···················· ···················· ···················· ····················
C 4489 hCoV-19/Sweden/20-08101/2020|EPI_ISL_454472|2020-0 ···················· ···················· ···················· ···················· ···················· ····················
C 4490 hCoV-19/Sweden/20-08102/2020|EPI_ISL_454473|2020-0 ···················· ···················· ···················· ···················· ···················· ····················
C 4491 hCoV-19/Sweden/20-08103/2020|EPI_ISL_454474|2020-0 ···················· ···················· ···················· ···················· ···················· ····················
C 4492 hCoV-19/Sweden/20-08104/2020|EPI_ISL_454475|2020-0 ···················· ···················· ···················· ···················· ···················· ····················
C 4493 hCoV-19/Sweden/20-08105/2020|EPI_ISL_454476|2020-0 ···················· ···················· ···················· ···················· ···················· ····················
C 4494 hCoV-19/Sweden/20-08106/2020|EPI_ISL_454477|2020-0 ···················· ···················· ···················· ···················· ···················· ····················
C 4495 hCoV-19/Sweden/20-08107/2020|EPI_ISL_454478|2020-0 ···················· ···················· ···················· ···················· ···················· ····················
C 4496 hCoV-19/Sweden/20-08108/2020|EPI_ISL_454479|2020-0 ···················· ···················· ···················· ···················· ···················· ····················
C 4497 hCoV-19/Sweden/20-08109/2020|EPI_ISL_454480|2020-0 ···················· ···················· ···················· ···················· ···················· ····················
C 4498 hCoV-19/Sweden/20-08110/2020|EPI_ISL_454481|2020-0 ···················· ···················· ···················· ···················· ···················· ····················
C 4499 hCoV-19/Sweden/20-08111/2020|EPI_ISL_454482|2020-0 ···················· ···················· ···················· ···················· ···················· ····················
C 4500 hCoV-19/Sweden/20-08112/2020|EPI_ISL_454483|2020-0 ···················· ···················· ···················· ···················· ···················· ····················
C 4501 hCoV-19/Sweden/20-08113/2020|EPI_ISL_454484|2020-0 ···················· ···················· ···················· ···················· ···················· ····················
C 4502 hCoV-19/Sweden/20-08114/2020|EPI_ISL_454485|2020-0 ···················· ···················· ···················· ···················· ···················· ····················
C 4503 hCoV-19/Sweden/20-08115/2020|EPI_ISL_454486|2020-0 ···················· ···················· ···················· ···················· ···················· ····················
C 4504 hCoV-19/Sweden/20-08116/2020|EPI_ISL_454487|2020-0 ···················· ···················· ···················· ···················· ···················· ····················
C 4505 hCoV-19/Sweden/20-08118/2020|EPI_ISL_454488|2020-0 ···················· ···················· ···················· ···················· ···················· ····················
C 4506 hCoV-19/Sweden/20-08119/2020|EPI_ISL_454489|2020-0 ···················· ···················· ···················· ···················· ···················· ····················
C 4507 hCoV-19/Sweden/20-08120/2020|EPI_ISL_454490|2020-0 ···················· ···················· ···················· ···················· ···················· ····················
C 4508 hCoV-19/Sweden/20-08121/2020|EPI_ISL_454491|2020-0 ···················· ···················· ···················· ···················· ···················· ····················
C 4509 hCoV-19/Sweden/20-08122/2020|EPI_ISL_454492|2020-0 ···················· ···················· ···················· ···················· ···················· ····················
C 4510 hCoV-19/Sweden/20-08123/2020|EPI_ISL_454493|2020-0 ···················· ···················· ···················· ···················· ···················· ····················
C 4511 hCoV-19/Sweden/20-08045/2020|EPI_ISL_454494|2020-0 ···················· ···················· ···················· ···················· ···················· ····················
C 4512 hCoV-19/Sweden/20-08047/2020|EPI_ISL_454495|2020-0 ···················· ···················· ···················· ···················· ···················· ····················
C 4513 hCoV-19/Sweden/20-08054/2020|EPI_ISL_454496|2020-0 ···················· ···················· ···················· ···················· ···················· ····················
C 4514 hCoV-19/Kazakhstan/16173/2020|EPI_ISL_454497|2020- ···················· ···················· ···················· ···················· ···················· ····················
C 4515 hCoV-19/Kazakhstan/16238/2020|EPI_ISL_454498|2020- ···················· ···················· ···················· ···················· ···················· ····················
C 4516 hCoV-19/Kazakhstan/16537/2020|EPI_ISL_454499|2020- ···················· ···················· ···················· ···················· ···················· ····················
C 4517 hCoV-19/Spain/VC-FISABIO-658/2020|EPI_ISL_455733|2 ···················· ···················· ···················· ···················· ···················· ····················
C 4518 hCoV-19/Spain/VC-FISABIO-h2647/2020|EPI_ISL_455735 ···················· ···················· ···················· ···················· ···················· ····················
C 4519 hCoV-19/Spain/VC-FISABIO-h2650/2020|EPI_ISL_455736 ···················· ···················· ···················· ···················· ···················· ··NNN··············· 1
C 4521 hCoV-19/India/OR-RMRC106/2020|EPI_ISL_455751|2020- ···················· ···················· ···················· ···················· ···················· ····················
C 4522 hCoV-19/India/OR-RMRC108/2020|EPI_ISL_455752|2020- ···················· ···················· ···················· ···················· ···················· ····················
C 4523 hCoV-19/India/OR-RMRC112/2020|EPI_ISL_455754|2020- ···················· ···················· ···················· ···················· ···················· ····················
C 4524 hCoV-19/India/OR-RMRC154/2020|EPI_ISL_455755|2020- ···················· ···················· ···················· ···················· ···················· ····················
C 4525 hCoV-19/India/OR-RMRC156/2020|EPI_ISL_455757|2020- ···················· ···················· ···················· ···················· ···················· ····················
C 4526 hCoV-19/India/OR-RMRC157/2020|EPI_ISL_455758|2020- ···················· ···················· ···················· ···················· ···················· ····················
C 4527 hCoV-19/India/OR-RMRC159/2020|EPI_ISL_455760|2020- ···················· ···················· ···················· ···················· ···················· ····················
C 4528 hCoV-19/India/OR-RMRC160/2020|EPI_ISL_455761|2020- ···················· ···················· ···················· ···················· ···················· ····················
C 4529 hCoV-19/India/OR-RMRC163/2020|EPI_ISL_455763|2020- ···················· ···················· ···················· ···················· ···················· ····················
C 4530 hCoV-19/India/OR-RMRC164/2020|EPI_ISL_455764|2020- ···················· ···················· ···················· ···················· ···················· ····················
C 4531 hCoV-19/India/OR-RMRC165/2020|EPI_ISL_455765|2020- ···················· ···················· ···················· ···················· ···················· ····················
C 4532 hCoV-19/India/OR-RMRC166/2020|EPI_ISL_455766|2020- ···················· ···················· ···················· ···················· ···················· ····················
C 4533 hCoV-19/India/OR-RMRC167/2020|EPI_ISL_455767|2020- ···················· ···················· ···················· ···················· ···················· ····················
C 4534 hCoV-19/India/OR-RMRC169/2020|EPI_ISL_455768|2020- ···················· ···················· ···················· ···················· ···················· ····················
C 4535 hCoV-19/India/OR-RMRC171/2020|EPI_ISL_455770|2020- ···················· ···················· ···················· ···················· ···················· ····················
C 4536 hCoV-19/India/OR-RMRC2/2020|EPI_ISL_455771|2020-03 ···················· ···················· ···················· ···················· ···················· ····················
C 4537 hCoV-19/India/OR-RMRC24/2020|EPI_ISL_455775|2020-0 ···················· ···················· ···················· ···················· ···················· ····················
C 4538 hCoV-19/India/OR-RMRC27/2020|EPI_ISL_455776|2020-0 ···················· ···················· ···················· ···················· ···················· ····················
C 4539 hCoV-19/India/OR-RMRC28/2020|EPI_ISL_455777|2020-0 ···················· ···················· ···················· ···················· ···················· ····················
C 4540 hCoV-19/India/OR-RMRC30/2020|EPI_ISL_455778|2020-0 ···················· ···················· ···················· ···················· ···················· ····················
C 4541 hCoV-19/India/OR-RMRC38/2020|EPI_ISL_455779|2020-0 ···················· ···················· ···················· ···················· ···················· ····················
C 4542 hCoV-19/India/OR-RMRC46/2020|EPI_ISL_455780|2020-0 ···················· ···················· ···················· ···················· ···················· ····················
C 4543 hCoV-19/India/OR-RMRC5/2020|EPI_ISL_455782|2020-04 ···················· ···················· ···················· ···················· ···················· ····················
C 4544 hCoV-19/India/OR-RMRC6/2020|EPI_ISL_455783|2020-04 ···················· ···················· ···················· ···················· ···················· ····················
C 4545 hCoV-19/India/OR-RMRC7/2020|EPI_ISL_455784|2020-04 ···················· ···················· ···················· ···················· ···················· ····················
C 4546 hCoV-19/India/OR-RMRC90/2020|EPI_ISL_455786|2020-0 ···················· ···················· ···················· ···················· ···················· ····················
C 4547 hCoV-19/India/OR-RMRC92/2020|EPI_ISL_455787|2020-0 ···················· ···················· ···················· ···················· ···················· ····················
C 4548 hCoV-19/Malaysia/IMR_WC119/2020|EPI_ISL_455790|202 ···················· ···················· ···················· ···················· ···················· ····················
C 4549 hCoV-19/Malaysia/IMR_WC413/2020|EPI_ISL_455791|202 ···················· ···················· ···················· ···················· ···················· ····················
C 4550 hCoV-19/Malaysia/IMR_WC458/2020|EPI_ISL_455792|202 ···················· ···················· ···················· ···················· ···················· ····················
C 4551 hCoV-19/Malaysia/IMR_WC1114/2020|EPI_ISL_455793|20 ···················· ···················· ···················· ···················· ···················· ····················

| | | | | | | | | |
|---|---|---|---|---|---|---|---|---|
| C | 4552 | hCoV-19/England/CAMB-1B0208/2020\|EPI_ISL_459271\|20 | ···················· | ···················· | ···················· | ···················· | ···················· | ···················· |
| C | 4553 | hCoV-19/England/CAMB-1AEF51/2020\|EPI_ISL_459274\|20 | ···················· | ···················· | ···················· | ···················· | ···················· | ···················· |
| C | 4554 | hCoV-19/England/CAMB-1B03C9/2020\|EPI_ISL_459284\|20 | ···················· | ···················· | ···················· | ···················· | ···················· | ···················· |
| C | 4555 | hCoV-19/England/CAMB-1AF679/2020\|EPI_ISL_459287\|20 | ···················· | ···················· | ···················· | ···················· | ···················· | ···················· |
| C | 4556 | hCoV-19/England/CAMB-1AE8DE/2020\|EPI_ISL_459289\|20 | ···················· | ···················· | ···················· | ···················· | ···················· | ···················· |
| C | 4557 | hCoV-19/England/CAMB-1B0271/2020\|EPI_ISL_459292\|20 | ···················· | ·A·················· | ···················· | ···················· | ···················· | ···················· |
| C | 4558 | hCoV-19/England/CAMB-1AF06C/2020\|EPI_ISL_459295\|20 | ···················· | ···················· | ···················· | ···················· | ···················· | ···················· |
| C | 4559 | hCoV-19/England/CAMB-1AF05D/2020\|EPI_ISL_459296\|20 | ···················· | ···················· | ···················· | ···················· | ···················· | ···················· |
| C | 4560 | hCoV-19/USA/MA-MGH-00514/2020\|EPI_ISL_460253\|2020- | ···················· | ···················· | ···················· | ···················· | ···················· | ···················· |
| C | 4561 | hCoV-19/USA/MA-MGH-00650/2020\|EPI_ISL_460254\|2020- | ···················· | ···················· | ···················· | ···················· | ···················· | ···················· |
| C | 4562 | hCoV-19/USA/MA-MGH-00299/2020\|EPI_ISL_460255\|2020- | ···················· | ···················· | ···················· | ···················· | ···················· | ···················· |
| C | 4563 | hCoV-19/USA/MA-MGH-00657/2020\|EPI_ISL_460256\|2020- | ···················· | ···················· | ···················· | ···················· | ···················· | ···················· |
| C | 4564 | hCoV-19/USA/MA-MGH-00688/2020\|EPI_ISL_460257\|2020- | ···················· | ···················· | ···················· | ···················· | ···················· | ···················· |
| C | 4565 | hCoV-19/USA/MA-MGH-00501/2020\|EPI_ISL_460258\|2020- | ···················· | ···················· | ···················· | ···················· | ···················· | ···················· |
| C | 4566 | hCoV-19/USA/MA-MGH-00156/2020\|EPI_ISL_460261\|2020- | ···················· | ···················· | ···················· | ···················· | ···················· | ···················· |
| C | 4567 | hCoV-19/USA/MA-MGH-00190/2020\|EPI_ISL_460263\|2020- | ···················· | ···················· | ···················· | ···················· | ···················· | ···················· |
| C | 4568 | hCoV-19/USA/MA-MGH-00049/2020\|EPI_ISL_460264\|2020- | ···················· | ···················· | ···················· | ···················· | ···················· | ···················· |
| C | 4569 | hCoV-19/USA/MA-MGH-00298/2020\|EPI_ISL_460265\|2020- | ···················· | ···················· | ···················· | ···················· | ···················· | ···················· |
| C | 4570 | hCoV-19/USA/MA-MGH-00421/2020\|EPI_ISL_460266\|2020- | ···················· | ···················· | ···················· | ···················· | ···················· | ···················· |
| C | 4571 | hCoV-19/USA/MA-MGH-00681/2020\|EPI_ISL_460267\|2020- | ···················· | ···················· | ···················· | ···················· | ···················· | ···················· |
| C | 4572 | hCoV-19/USA/MA-MGH-00699/2020\|EPI_ISL_460268\|2020- | ···················· | ···················· | ···················· | ···················· | ···················· | ···················· |
| C | 4573 | hCoV-19/USA/MA-MGH-00093/2020\|EPI_ISL_460269\|2020- | ···················· | ···················· | ···················· | ···················· | ···················· | ···················· |
| C | 4574 | hCoV-19/USA/MA-MGH-00604/2020\|EPI_ISL_460270\|2020- | ···················· | ···················· | ···················· | ···················· | ···················· | ···················· |
| C | 4575 | hCoV-19/USA/MA-MGH-00014/2020\|EPI_ISL_460271\|2020- | ···················· | ···················· | ···················· | ···················· | ···················· | ···················· |
| C | 4576 | hCoV-19/USA/MA-MGH-00458/2020\|EPI_ISL_460272\|2020- | ···················· | ···················· | ···················· | ···················· | ···················· | ···················· |
| C | 4577 | hCoV-19/USA/MA-MGH-00580/2020\|EPI_ISL_460273\|2020- | ···················· | ···················· | ···················· | ···················· | ···················· | ···················· |
| C | 4578 | hCoV-19/USA/MA-MGH-00294/2020\|EPI_ISL_460274\|2020- | ···················· | ···················· | ···················· | ···················· | ···················· | ···················· |
| C | 4579 | hCoV-19/USA/MA-MGH-00538/2020\|EPI_ISL_460275\|2020- | ···················· | ···················· | ···················· | ···················· | ···················· | ···················· |
| C | 4580 | hCoV-19/USA/MA-MGH-00583/2020\|EPI_ISL_460276\|2020- | ···················· | ···················· | ···················· | ···················· | ···················· | ···················· |
| C | 4581 | hCoV-19/USA/MA-MGH-00716/2020\|EPI_ISL_460277\|2020- | ···················· | ···················· | ···················· | ···················· | ···················· | ···················· |
| C | 4582 | hCoV-19/USA/MA-MGH-00715/2020\|EPI_ISL_460278\|2020- | ···················· | ···················· | ···················· | ···················· | ···················· | ···················· |
| C | 4583 | hCoV-19/USA/MA-MGH-00597/2020\|EPI_ISL_460279\|2020- | ···················· | ···················· | ···················· | ···················· | ···················· | ···················· |
| C | 4584 | hCoV-19/USA/MA-MGH-00209/2020\|EPI_ISL_460280\|2020- | ···················· | ···················· | ···················· | ···················· | ···················· | ···················· |
| C | 4585 | hCoV-19/USA/MA-MGH-00094/2020\|EPI_ISL_460281\|2020- | ···················· | ···················· | ···················· | ···················· | ···················· | ···················· |
| C | 4586 | hCoV-19/USA/MA-MGH-00645/2020\|EPI_ISL_460282\|2020- | ···················· | ···················· | ···················· | ···················· | ···················· | ···················· |
| C | 4587 | hCoV-19/USA/MA-MGH-00170/2020\|EPI_ISL_460283\|2020- | ···················· | ···················· | ···················· | ···················· | ···················· | ···················· |
| C | 4588 | hCoV-19/USA/MA-MGH-00453/2020\|EPI_ISL_460284\|2020- | ···················· | ···················· | ···················· | ···················· | ···················· | ···················· |
| C | 4589 | hCoV-19/USA/MA-MGH-00566/2020\|EPI_ISL_460285\|2020- | ···················· | ···················· | ···················· | ···················· | ···················· | ···················· |
| C | 4590 | hCoV-19/USA/MA-MGH-00484/2020\|EPI_ISL_460286\|2020- | ···················· | ···················· | ···················· | ···················· | ···················· | ···················· |
| C | 4591 | hCoV-19/USA/MA-MGH-00203/2020\|EPI_ISL_460288\|2020- | ···················· | ···················· | ···················· | ···················· | ···················· | ···················· |
| C | 4592 | hCoV-19/USA/MA-MGH-00622/2020\|EPI_ISL_460289\|2020- | ···················· | ···················· | ···················· | ···················· | ···················· | ···················· |
| C | 4593 | hCoV-19/USA/MA-MGH-00166/2020\|EPI_ISL_460290\|2020- | ···················· | ···················· | ···················· | ···················· | ···················· | ···················· |
| C | 4594 | hCoV-19/USA/MA-MGH-00048/2020\|EPI_ISL_460291\|2020- | ···················· | ···················· | ···················· | ···················· | ···················· | ···················· |
| C | 4595 | hCoV-19/USA/MA-MGH-00052/2020\|EPI_ISL_460292\|2020- | ···················· | ···················· | ···················· | ···················· | ···················· | ···················· |
| C | 4596 | hCoV-19/USA/MA-MGH-00705/2020\|EPI_ISL_460293\|2020- | ···················· | ···················· | ···················· | ···················· | ···················· | ···················· |
| C | 4597 | hCoV-19/USA/MA-MGH-00265/2020\|EPI_ISL_460294\|2020- | ···················· | ···················· | ···················· | ···················· | ···················· | ···················· |
| C | 4598 | hCoV-19/USA/MA-MGH-00595/2020\|EPI_ISL_460295\|2020- | ···················· | ···················· | ···················· | ···················· | ···················· | ···················· |
| C | 4599 | hCoV-19/USA/MA-MGH-00668/2020\|EPI_ISL_460296\|2020- | ···················· | ···················· | ···················· | ···················· | ···················· | ···················· |
| C | 4600 | hCoV-19/England/CAMB-1B3E3E/2020\|EPI_ISL_461586\|20 | ···················· | ···················· | ···················· | ···················· | ···················· | ···················· |
| C | 4601 | hCoV-19/England/CAMB-1B3E7A/2020\|EPI_ISL_461587\|20 | ···················· | ···················· | ···················· | ···················· | ···················· | ···················· |
| C | 4602 | hCoV-19/Scotland/CVR2920/2020\|EPI_ISL_461589\|2020- | ···················· | ···················· | ···················· | ···················· | ···················· | ···················· |
| C | 4604 | hCoV-19/Scotland/CVR2929/2020\|EPI_ISL_461595\|2020- | ···················· | ···················· | ···················· | ···················· | ···················· | ···················· |
| C | 4605 | hCoV-19/Scotland/CVR2931/2020\|EPI_ISL_461596\|2020- | ···················· | ···················· | ···················· | ···················· | ···················· | ···················· |
| C | 4606 | hCoV-19/Scotland/CVR2933/2020\|EPI_ISL_461597\|2020- | ···················· | ···················· | ···················· | ···················· | ···················· | ···················· |
| C | 4607 | hCoV-19/Scotland/CVR2934/2020\|EPI_ISL_461598\|2020- | ···················· | ··············C····· | ···················· | ···················· | ···················· | ···················· |
| C | 4609 | hCoV-19/Israel/CVL-n3120/2020\|EPI_ISL_447251\|2020- | ···················· | ···················· | ···················· | ···················· | ···················· | ···················· |
| C | 4610 | hCoV-19/Taiwan/TSGH-26/2020\|EPI_ISL_447252\|2020-02 | ···················· | ···················· | ···················· | ···················· | ···················· | ···················· |
| C | 4611 | hCoV-19/Taiwan/TSGH-27/2020\|EPI_ISL_447253\|2020-03 | ···················· | ···················· | ···················· | ···················· | ···················· | ···················· |
| C | 4612 | hCoV-19/Taiwan/TSGH-29/2020\|EPI_ISL_447255\|2020-03 | ···················· | ···················· | ···················· | ···················· | ···················· | ···················· |
| C | 4613 | hCoV-19/Taiwan/TSGH-31/2020\|EPI_ISL_447257\|2020-03 | ···················· | ···················· | ···················· | ···················· | ···················· | ···················· |
| C | 4614 | hCoV-19/Israel/990333193/2020\|EPI_ISL_447260\|2020- | ···················· | ···················· | ···················· | ···················· | ···················· | ···················· |
| C | 4615 | hCoV-19/Israel/990307712/2020\|EPI_ISL_447263\|2020- | ···················· | ···················· | ···················· | ···················· | ···················· | ···················· |
| C | 4616 | hCoV-19/Israel/990300860/2020\|EPI_ISL_447264\|2020- | ···················· | ···················· | ···················· | ···················· | ···················· | ···················· |
| C | 4617 | hCoV-19/Israel/990300724/2020\|EPI_ISL_447265\|2020- | ···················· | ···················· | ···················· | ···················· | ···················· | ···················· |
| C | 4618 | hCoV-19/Israel/990300681/2020\|EPI_ISL_447267\|2020- | ···················· | ···················· | ···················· | ···················· | ···················· | ···················· |
| C | 4619 | hCoV-19/Israel/990059252/2020\|EPI_ISL_447268\|2020- | ···················· | ···················· | ···················· | ···················· | ···················· | ···················· |
| C | 4620 | hCoV-19/Israel/990059251/2020\|EPI_ISL_447269\|2020- | ···················· | ···················· | ···················· | ···················· | ···················· | ···················· |
| C | 4621 | hCoV-19/Israel/990059238/2020\|EPI_ISL_447271\|2020- | ···················· | ···················· | ···················· | ···················· | ···················· | ···················· |
| C | 4622 | hCoV-19/Israel/990059237/2020\|EPI_ISL_447272\|2020- | ···················· | ···················· | ···················· | ···················· | ···················· | ···················· |
| C | 4623 | hCoV-19/Israel/990059233/2020\|EPI_ISL_447273\|2020- | ···················· | ···················· | ···················· | ···················· | ···················· | ···················· |
| C | 4624 | hCoV-19/Israel/990059232/2020\|EPI_ISL_447274\|2020- | ···················· | ···················· | ···················· | ···················· | ···················· | ·········NN········· |
| C | 4625 | hCoV-19/Israel/990059231/2020\|EPI_ISL_447275\|2020- | ···················· | ···················· | ···················· | ···················· | ···················· | ···················· |
| C | 4626 | hCoV-19/Israel/990059217/2020\|EPI_ISL_447277\|2020- | ···················· | ···················· | ···················· | ···················· | ···················· | ···················· |
| C | 4627 | hCoV-19/Israel/990059203/2020\|EPI_ISL_447279\|2020- | ···················· | ···················· | ···················· | ···················· | ···················· | ···················· |
| C | 4628 | hCoV-19/Israel/990059202/2020\|EPI_ISL_447280\|2020- | ···················· | ···················· | ···················· | ···················· | ···················· | ·········N·········· |
| C | 4629 | hCoV-19/Israel/701002792/2020\|EPI_ISL_447281\|2020- | ···················· | ···················· | ···················· | ···················· | ···················· | ···················· |
| C | 4630 | hCoV-19/Israel/701002786/2020\|EPI_ISL_447282\|2020- | ···················· | ···················· | ···················· | ···················· | ···················· | ···················· |
| C | 4631 | hCoV-19/Israel/701002768/2020\|EPI_ISL_447283\|2020- | ···················· | ···················· | ···················· | ···················· | ···················· | ···················· |

| | | | | | | | | |
|---|---|---|---|---|---|---|---|---|
| C | 4633 | hCoV-19/Israel/701002681/2020\|EPI_ISL_447285\|2020- | ···················· | ···················· | ····················· | ···················· | ···················· | ···················· |
| C | 4634 | hCoV-19/Israel/701002666/2020\|EPI_ISL_447286\|2020- | ···················· | ···················· | ····················· | ···················· | ···················· | ···················· |
| C | 4635 | hCoV-19/Israel/701002591/2020\|EPI_ISL_447287\|2020- | ···················· | ···················· | ····················· | ···················· | ···················· | ···················· |
| C | 4636 | hCoV-19/Israel/701002561/2020\|EPI_ISL_447288\|2020- | ···················· | ···················· | ····················· | ···················· | ···················· | ···················· |
| C | 4637 | hCoV-19/Israel/701002556/2020\|EPI_ISL_447289\|2020- | ···················· | ···················· | ····················· | ···················· | ···················· | ···················· |
| C | 4638 | hCoV-19/England/OXON-ACDB4/2020\|EPI_ISL_448538\|202 | ···················· | ·············A······ | ····················· | ···················· | ···················· | ···················· |
| C | 4639 | hCoV-19/England/OXON-ACE2A/2020\|EPI_ISL_448540\|202 | ···················· | ···················· | ····················· | ···················· | ···················· | ···················· |
| C | 4640 | hCoV-19/England/OXON-ACE57/2020\|EPI_ISL_448541\|202 | ···················· | ···················· | ····················· | ···················· | ···················· | ···················· |
| C | 4641 | hCoV-19/England/OXON-ACE75/2020\|EPI_ISL_448542\|202 | ···················· | ···················· | ····················· | ···················· | ···················· | ···················· |
| C | 4642 | hCoV-19/England/OXON-ACEC0/2020\|EPI_ISL_448545\|202 | ···················· | ···················· | ····················· | ···················· | ···················· | ···················· |
| C | 4643 | hCoV-19/England/OXON-ACEEE/2020\|EPI_ISL_448546\|202 | ···················· | ···················· | ····················· | ···················· | ···················· | ···················· |
| C | 4644 | hCoV-19/England/OXON-ACEFD/2020\|EPI_ISL_448547\|202 | ···················· | ···················· | ····················· | ···················· | ···················· | ···················· |
| C | 4645 | hCoV-19/England/OXON-ACF54/2020\|EPI_ISL_448548\|202 | ···················· | ···················· | ····················· | ···················· | ···················· | ···················· |
| C | 4646 | hCoV-19/England/OXON-ACFAF/2020\|EPI_ISL_448549\|202 | ···················· | ···················· | ····················· | ···················· | ···················· | ···················· |
| C | 4647 | hCoV-19/England/OXON-ACFDC/2020\|EPI_ISL_448550\|202 | ···················· | ···················· | ····················· | ···················· | ···················· | ···················· |
| C | 4648 | hCoV-19/England/OXON-ACFEB/2020\|EPI_ISL_448551\|202 | ···················· | ·A·················· | ····················· | ···················· | ···················· | ···················· |
| C | 4649 | hCoV-19/England/OXON-ACFFA/2020\|EPI_ISL_448552\|202 | ···················· | ···················· | ····················· | ···················· | ···················· | ···················· |
| C | 4650 | hCoV-19/England/OXON-AD005/2020\|EPI_ISL_448553\|202 | ···················· | ···················· | ····················· | ···················· | ···················· | ···················· |
| C | 4651 | hCoV-19/England/OXON-AD041/2020\|EPI_ISL_448555\|202 | ···················· | ·············A······ | ····················· | ···················· | ···················· | ···················· |
| C | 4652 | hCoV-19/England/OXON-AD050/2020\|EPI_ISL_448556\|202 | ···················· | ···················· | ····················· | ···················· | ···················· | ···················· |
| C | 4653 | hCoV-19/England/OXON-AD08D/2020\|EPI_ISL_448557\|202 | ···················· | ···················· | ····················· | ···················· | ···················· | ···················· |
| C | 4654 | hCoV-19/England/OXON-AD0AB/2020\|EPI_ISL_448559\|202 | ···················· | ···················· | ····················· | ···················· | ···················· | ···················· |
| C | 4655 | hCoV-19/England/OXON-AD0C9/2020\|EPI_ISL_448561\|202 | ···················· | ·A·················· | ····················· | ···················· | ···················· | ···················· |
| C | 4656 | hCoV-19/England/OXON-AD0F6/2020\|EPI_ISL_448563\|202 | ···················· | ···················· | ····················· | ···················· | ···················· | ···················· |
| C | 4657 | hCoV-19/England/OXON-AD102/2020\|EPI_ISL_448564\|202 | ···················· | ···················· | ····················· | ···················· | ···················· | ···················· |
| C | 4658 | hCoV-19/England/OXON-AD120/2020\|EPI_ISL_448565\|202 | ···················· | ···················· | ····················· | ···················· | ···················· | ···················· |
| C | 4659 | hCoV-19/England/OXON-AD13F/2020\|EPI_ISL_448566\|202 | ···················· | ···················· | ····················· | ···················· | ···················· | ···················· |
| C | 4660 | hCoV-19/England/OXON-AD15D/2020\|EPI_ISL_448567\|202 | ···················· | ···················· | ····················· | ···················· | ···················· | ···················· |
| C | 4661 | hCoV-19/England/OXON-AD1B7/2020\|EPI_ISL_448568\|202 | ···················· | ···················· | ····················· | ···················· | ···················· | ···················· |
| C | 4662 | hCoV-19/England/OXON-AD21E/2020\|EPI_ISL_448570\|202 | ···················· | ···················· | ····················· | ···················· | ···················· | ···················· |
| C | 4663 | hCoV-19/England/OXON-AD23C/2020\|EPI_ISL_448571\|202 | ···················· | ···················· | ····················· | ···················· | ···················· | ···················· |
| C | 4664 | hCoV-19/England/OXON-AD25A/2020\|EPI_ISL_448572\|202 | ···················· | ···················· | ····················· | ···················· | ···················· | ···················· |
| C | 4665 | hCoV-19/England/OXON-AD296/2020\|EPI_ISL_448573\|202 | ···················· | ···················· | ····················· | ···················· | ···················· | ···················· |
| C | 4666 | hCoV-19/England/OXON-AD31B/2020\|EPI_ISL_448575\|202 | ···················· | ···················· | ····················· | ···················· | ···················· | ···················· |
| C | 4667 | hCoV-19/England/OXON-AD375/2020\|EPI_ISL_448577\|202 | ···················· | ···················· | ····················· | ···················· | ···················· | ···················· |
| C | 4668 | hCoV-19/England/OXON-AD3DF/2020\|EPI_ISL_448579\|202 | ···················· | ···················· | ····················· | ···················· | ···················· | ···················· |
| C | 4669 | hCoV-19/England/OXON-AD454/2020\|EPI_ISL_448580\|202 | ···················· | ···················· | ····················· | ···················· | ···················· | ···················· |
| C | 4670 | hCoV-19/England/OXON-AD4BE/2020\|EPI_ISL_448582\|202 | ···················· | ···················· | ····················· | ···················· | ···················· | ···················· |
| C | 4671 | hCoV-19/England/OXON-AD4FA/2020\|EPI_ISL_448584\|202 | ···················· | ···················· | ····················· | ···················· | ···················· | ···················· |
| C | 4672 | hCoV-19/England/OXON-AD515/2020\|EPI_ISL_448585\|202 | ···················· | ···················· | ····················· | ···················· | ···················· | ···················· |
| C | 4673 | hCoV-19/England/OXON-AD67C/2020\|EPI_ISL_448587\|202 | ···················· | ···················· | ····················· | ···················· | ···················· | ···················· |
| C | 4674 | hCoV-19/England/OXON-AD6B8/2020\|EPI_ISL_448588\|202 | ···················· | ···················· | ····················· | ···················· | ···················· | ···················· |
| C | 4675 | hCoV-19/England/OXON-AD6F4/2020\|EPI_ISL_448590\|202 | ···················· | ···················· | ····················· | ···················· | ···················· | ···················· |
| C | 4676 | hCoV-19/England/OXON-AD71F/2020\|EPI_ISL_448591\|202 | ···················· | ···················· | ····················· | ···················· | ···················· | ···················· |
| C | 4677 | hCoV-19/England/OXON-AD73D/2020\|EPI_ISL_448592\|202 | ···················· | ···················· | ····················· | ···················· | ···················· | ···················· |
| C | 4678 | hCoV-19/England/OXON-AD75B/2020\|EPI_ISL_448593\|202 | ···················· | ···················· | ····················· | ···················· | ···················· | ···················· |
| C | 4679 | hCoV-19/England/OXON-AD779/2020\|EPI_ISL_448594\|202 | ···················· | ···················· | ····················· | ···················· | ···················· | ···················· |
| C | 4680 | hCoV-19/England/OXON-AD7B5/2020\|EPI_ISL_448595\|202 | ···················· | ···················· | ····················· | ···················· | ···················· | ···················· |
| C | 4681 | hCoV-19/England/OXON-AD7D3/2020\|EPI_ISL_448596\|202 | ···················· | ···················· | ····················· | ···················· | ···················· | ···················· |
| C | 4682 | hCoV-19/England/OXON-AD81C/2020\|EPI_ISL_448597\|202 | ···················· | ···················· | ····················· | ···················· | ···················· | ···················· |
| C | 4683 | hCoV-19/England/OXON-AD83A/2020\|EPI_ISL_448598\|202 | ···················· | ···················· | ····················· | ···················· | ···················· | ···················· |
| C | 4684 | hCoV-19/England/OXON-AD858/2020\|EPI_ISL_448599\|202 | ···················· | ···················· | ····················· | ···················· | ···················· | ···················· |
| C | 4685 | hCoV-19/USA/UT-00361/2020\|EPI_ISL_449810\|2020-03-3 | ···················· | ···················· | ····················· | ···················· | ···················· | ···················· |
| C | 4686 | hCoV-19/USA/UT-01314/2020\|EPI_ISL_449812\|2020-04-0 | ···················· | ···················· | ····················· | ···················· | ···················· | ···················· |
| C | 4687 | hCoV-19/USA/WA-S699/2020\|EPI_ISL_449869\|2020-04-11 | ···················· | ···················· | ····················· | ···················· | ···················· | ···················· |
| C | 4688 | hCoV-19/USA/WA-S700/2020\|EPI_ISL_449870\|2020-04-09 | ···················· | ···················· | ····················· | ···················· | ···················· | ···················· |
| C | 4689 | hCoV-19/USA/WA-S701/2020\|EPI_ISL_449871\|2020-04-10 | ···················· | ···················· | ····················· | ···················· | ···················· | ···················· |
| C | 4690 | hCoV-19/USA/WA-S702/2020\|EPI_ISL_449872\|2020-04-10 | ···················· | ···················· | ····················· | ···················· | ···················· | ···················· |
| C | 4691 | hCoV-19/USA/WA-S703/2020\|EPI_ISL_449873\|2020-04-10 | ···················· | ···················· | ····················· | ···················· | ···················· | ···················· |
| C | 4692 | hCoV-19/USA/WA-S704/2020\|EPI_ISL_449874\|2020-04-09 | ···················· | ···················· | ····················· | ···················· | ···················· | ···················· |
| C | 4693 | hCoV-19/USA/WA-S705/2020\|EPI_ISL_449875\|2020-04-09 | ···················· | ···················· | ····················· | ···················· | ···················· | ···················· |
| C | 4694 | hCoV-19/USA/WA-S706/2020\|EPI_ISL_449876\|2020-04-08 | ···················· | ···················· | ····················· | ···················· | ···················· | ···················· |
| C | 4695 | hCoV-19/USA/WA-S707/2020\|EPI_ISL_449877\|2020-04-07 | ···················· | ···················· | ····················· | ···················· | ···················· | ···················· |
| C | 4696 | hCoV-19/USA/WA-S708/2020\|EPI_ISL_449878\|2020-04-09 | ···················· | ···················· | ····················· | ···················· | ···················· | ···················· |
| C | 4697 | hCoV-19/USA/WA-S709/2020\|EPI_ISL_449879\|2020-04-09 | ···················· | ···················· | ····················· | ···················· | ···················· | ···················· |
| C | 4698 | hCoV-19/USA/WA-S710/2020\|EPI_ISL_449880\|2020-04-07 | ···················· | ···················· | ····················· | ···················· | ···················· | ···················· |
| C | 4699 | hCoV-19/USA/WA-S711/2020\|EPI_ISL_449881\|2020-04-08 | ···················· | ···················· | ····················· | ···················· | ···················· | ···················· |
| C | 4700 | hCoV-19/USA/WA-S712/2020\|EPI_ISL_449882\|2020-04-07 | ···················· | ···················· | ····················· | ···················· | ···················· | ···················· |
| C | 4701 | hCoV-19/USA/WA-S713/2020\|EPI_ISL_449883\|2020-04-08 | ···················· | ···················· | ····················· | ···················· | ···················· | ···················· |
| C | 4702 | hCoV-19/USA/WA-S714/2020\|EPI_ISL_449884\|2020-04-07 | ···················· | ···················· | ····················· | ···················· | ···················· | ···················· |
| C | 4703 | hCoV-19/USA/WA-S715/2020\|EPI_ISL_449885\|2020-04-08 | ···················· | ···················· | ····················· | ···················· | ···················· | ···················· |
| C | 4704 | hCoV-19/USA/WA-S716/2020\|EPI_ISL_449886\|2020-04-11 | ···················· | ···················· | ····················· | ···················· | ···················· | ···················· |
| C | 4705 | hCoV-19/USA/WA-S717/2020\|EPI_ISL_449887\|2020-04-11 | ···················· | ···················· | ····················· | ···················· | ···················· | ···················· |
| C | 4706 | hCoV-19/USA/WA-S718/2020\|EPI_ISL_449888\|2020-04-11 | ···················· | ···················· | ····················· | ···················· | ···················· | ···················· |
| C | 4707 | hCoV-19/USA/WA-S719/2020\|EPI_ISL_449889\|2020-04-11 | ···················· | ···················· | ····················· | ···················· | ···················· | ···················· |
| C | 4708 | hCoV-19/USA/WA-S720/2020\|EPI_ISL_449890\|2020-04-12 | ···················· | ···················· | ····················· | ···················· | ···················· | ···················· |
| C | 4709 | hCoV-19/USA/WA-S721/2020\|EPI_ISL_449891\|2020-04-12 | ···················· | ···················· | ····················· | ···················· | ···················· | ···················· |
| C | 4710 | hCoV-19/USA/WA-S722/2020\|EPI_ISL_449892\|2020-04-12 | ···················· | ···················· | ····················· | ···················· | ···················· | ···················· |

```
C    4711 hCoV-19/USA/WA-S724/2020|EPI_ISL_449894|2020-04-10   ····················  ····················  ·····················  ····················  ····················  ····················
C    4712 hCoV-19/USA/WA-S725/2020|EPI_ISL_449895|2020-04-10   ····················  ····················  ·····················  ····················  ····················  ····················
C    4713 hCoV-19/USA/WA-S726/2020|EPI_ISL_449896|2020-04-11   ····················  ····················  ·····················  ····················  ····················  ····················
C    4714 hCoV-19/USA/WA-S727/2020|EPI_ISL_449897|2020-04-10   ····················  ····················  ·····················  ····················  ····················  ····················
C    4715 hCoV-19/USA/WA-S728/2020|EPI_ISL_449898|2020-04-10   ····················  ····················  ·····················  ····················  ····················  ····················
C    4716 hCoV-19/USA/WA-S729/2020|EPI_ISL_449899|2020-04-11   ·····T··············  ····················  ·····················  ····················  ····················  ····················
C    4717 hCoV-19/USA/AR-CDC-2072/2020|EPI_ISL_450800|2020-0   ····················  ····················  ·····················  ····················  ····················  ····················
C    4718 hCoV-19/USA/FL-BPHL-0116/2020|EPI_ISL_450850|2020-   ····················  ····················  ·····················  ····················  ····················  ····················
C    4719 hCoV-19/Brazil/AP-IEC-161167/2020|EPI_ISL_450873|2   ····················  ····················  ·····················  ····················  ····················  ····················
C    4720 hCoV-19/Brazil/PA-IEC-161548/2020|EPI_ISL_450874|2   ····················  ····················  ·····················  ····················  ····················  ····················
C    4721 hCoV-19/USA/PA-MGEL135/2020|EPI_ISL_454400|2020      ····················  ····················  ·····················  ····················  ····················  ····················
C    4722 hCoV-19/USA/PA-MGEL149/2020|EPI_ISL_454407|2020-04   ····················  ····················  ·····················  ····················  ····················  ····················
C    4723 hCoV-19/USA/PA-MGEL154/2020|EPI_ISL_454410|2020-04   ····················  ····················  ·····················  ····················  ····················  ····················
C    4724 hCoV-19/USA/PA-MGEL155/2020|EPI_ISL_454411|2020-04   ····················  ····················  ·····················  ····················  ····················  ····················
C    4725 hCoV-19/Vietnam/VNHN_2406/2020|EPI_ISL_455700|2020   ····················  ····················  ·····················  ····················  ····················  ····················
C    4726 hCoV-19/Vietnam/VNHN_1713/2020|EPI_ISL_455701|2020   ····················  ····················  ·····················  ····················  ····················  ····················
C    4727 hCoV-19/Vietnam/VNHN_3096/2020|EPI_ISL_455702|2020   ····················  ····················  ·····················  ····················  ····················  ····················
C    4728 hCoV-19/Vietnam/VNHN_3085/2020|EPI_ISL_455703|2020   ····················  ····················  ·····················  ····················  ····················  ····················
C    4729 hCoV-19/Vietnam/VNHN_3916/2020|EPI_ISL_455704|2020   ····················  ····················  ·····················  ····················  ····················  ····················
C    4730 hCoV-19/Vietnam/VNHN_3913/2020|EPI_ISL_455705|2020   ····················  ····················  ·····················  ····················  ····················  ····················
C    4731 hCoV-19/Vietnam/VNHN_3629/2020|EPI_ISL_455706|2020   ····················  ····················  ·····················  ····················  ····················  ····················
C    4732 hCoV-19/Vietnam/VNHN_4189/2020|EPI_ISL_455707|2020   ····················  ····················  ·····················  ····················  ····················  ····················
C    4733 hCoV-19/Vietnam/VNHN_4864/2020|EPI_ISL_455708|2020   ····················  ····················  ·····················  ····················  ····················  ····················
C    4734 hCoV-19/Vietnam/VNHN_4851/2020|EPI_ISL_455709|2020   ····················  ····················  ·····················  ····················  ····················  ····················
C    4735 hCoV-19/Vietnam/VNHN_4868/2020|EPI_ISL_455710|2020   ····················  ····················  ·····················  ····················  ····················  ····················
C    4736 hCoV-19/Vietnam/VNHN_4806/2020|EPI_ISL_455711|2020   ····················  ····················  ·····················  ····················  ····················  ····················
C    4737 hCoV-19/Vietnam/VNHN_4875/2020|EPI_ISL_455712|2020   ····················  ····················  ·····················  ····················  ····················  ····················
C    4738 hCoV-19/Vietnam/VNHN_301/2020|EPI_ISL_455714|2020-   ····················  ····················  ·····················  ····················  ····················  ····················
C    4739 hCoV-19/Vietnam/VNHN_4958/2020|EPI_ISL_455718|2020   ····················  ····················  ·····················  ····················  ····················  ····················
C    4740 hCoV-19/Spain/VC-FISABIO-661/2020|EPI_ISL_455727|2   ····················  ····················  ·····················  ····················  ····················  ····················
C    4742 hCoV-19/Spain/VC-FISABIO-650/2020|EPI_ISL_455741|2   ····················  ····················  ·····················  ····················  ····················  ··NNN···············   1
C    4744 hCoV-19/England/CAMB-1B1155/2020|EPI_ISL_459204|20   ····················  ····················  ·····················  ····················  ····················  ····················
C    4745 hCoV-19/England/CAMB-1B12F8/2020|EPI_ISL_459207|20   ····················  ····················  ·····················  ····················  ····················  ····················
C    4746 hCoV-19/England/CAMB-1AFC59/2020|EPI_ISL_459210|20   ····················  ····················  ·····················  ····················  ····················  ····················
C    4747 hCoV-19/England/CAMB-1AFC2C/2020|EPI_ISL_459211|20   ····················  ····················  ·····················  ····················  ····················  ····················
C    4748 hCoV-19/England/CAMB-1AFBC5/2020|EPI_ISL_459219|20   ····················  ····················  ·····················  ····················  ····················  ····················
C    4749 hCoV-19/England/CAMB-1B1119/2020|EPI_ISL_459227|20   ····················  ····················  ·····················  ····················  ····················  ····················
C    4750 hCoV-19/England/CAMB-1B095E/2020|EPI_ISL_459235|20   ····················  ····················  ·····················  ····················  ····················  ····················
C    4751 hCoV-19/England/CAMB-1AF697/2020|EPI_ISL_459240|20   ····················  ····················  ·····················  ····················  ····················  ····················
C    4752 hCoV-19/England/CAMB-1AF925/2020|EPI_ISL_459251|20   ····················  ····················  ·····················  ····················  ····················  ····················
C    4753 hCoV-19/England/CAMB-1AEF8E/2020|EPI_ISL_459252|20   ····················  ····················  ·····················  ····················  ····················  ····················
C    4754 hCoV-19/England/CAMB-1AFA13/2020|EPI_ISL_459255|20   ····················  ····················  ·····················  ····················  ····················  ····················
C    4755 hCoV-19/England/CAMB-1AF12D/2020|EPI_ISL_459257|20   ····················  ····················  ·····················  ····················  ····················  ····················
C    4756 hCoV-19/England/CAMB-1B0244/2020|EPI_ISL_459259|20   ····················  ····················  ·····················  ····················  ····················  ····················
C    4757 hCoV-19/England/CAMB-1AF10F/2020|EPI_ISL_459262|20   ····················  ····················  ·····················  ····················  ····················  ····················
C    4758 hCoV-19/England/CAMB-1B02DB/2020|EPI_ISL_459263|20   ····················  ····················  ·····················  ····················  ····················  ····················
C    4759 hCoV-19/England/CAMB-1AF521/2020|EPI_ISL_459264|20   ····················  ····················  ·····················  ····················  ····················  ····················
C    4760 hCoV-19/England/CAMB-1AEFCA/2020|EPI_ISL_459265|20   ····················  ····················  ·····················  ····················  ····················  ····················
C    4761 hCoV-19/England/CAMB-1AE8B0/2020|EPI_ISL_459266|20   ····················  ····················  ·····················  ····················  ····················  ····················
C    4762 hCoV-19/England/CAMB-1AF65B/2020|EPI_ISL_459269|20   ····················  ····················  ·····················  ····················  ····················  ····················
C    4763 hCoV-19/USA/MA-MGH-00473/2020|EPI_ISL_460201|2020-   ····················  ····················  ·····················  ····················  ····················  ····················
C    4764 hCoV-19/USA/MA-MGH-00076/2020|EPI_ISL_460202|2020-   ····················  ····················  ·····················  ····················  ····················  ····················
C    4765 hCoV-19/USA/MA-MGH-00195/2020|EPI_ISL_460203|2020-   ····················  ····················  ·····················  ····················  ····················  ····················
C    4766 hCoV-19/USA/MA-MGH-00486/2020|EPI_ISL_460204|2020-   ····················  ····················  ·····················  ····················  ····················  ····················
C    4767 hCoV-19/USA/MA-MGH-00614/2020|EPI_ISL_460205|2020-   ····················  ····················  ·····················  ····················  ····················  ····················
C    4768 hCoV-19/USA/MA-MGH-00032/2020|EPI_ISL_460206|2020-   ····················  ····················  ·····················  ····················  ····················  ····················
C    4769 hCoV-19/USA/MA-MGH-00442/2020|EPI_ISL_460207|2020-   ····················  ····················  ·····················  ····················  ····················  ····················
C    4770 hCoV-19/USA/MA-MGH-00477/2020|EPI_ISL_460208|2020-   ····················  ····················  ·····················  ····················  ····················  ····················
C    4771 hCoV-19/USA/MA-MGH-00177/2020|EPI_ISL_460209|2020-   ····················  ····················  ·····················  ····················  ····················  ····················
C    4772 hCoV-19/USA/MA-MGH-00280/2020|EPI_ISL_460210|2020-   ····················  ····················  ·····················  ····················  ····················  ····················
C    4773 hCoV-19/USA/MA-MGH-00427/2020|EPI_ISL_460212|2020-   ····················  ····················  ·····················  ····················  ····················  ····················
C    4774 hCoV-19/USA/MA-MGH-00247/2020|EPI_ISL_460213|2020-   ····················  ····················  ·····················  ····················  ····················  ····················
C    4775 hCoV-19/USA/MA-MGH-00063/2020|EPI_ISL_460214|2020-   ····················  ····················  ·····················  ····················  ····················  ····················
C    4776 hCoV-19/USA/MA-MGH-00693/2020|EPI_ISL_460215|2020-   ····················  ····················  ·····················  ····················  ····················  ····················
C    4777 hCoV-19/USA/MA-MGH-00541/2020|EPI_ISL_460216|2020-   ····················  ····················  ·····················  ····················  ····················  ····················
C    4778 hCoV-19/USA/MA-MGH-00659/2020|EPI_ISL_460217|2020-   ····················  ····················  ·····················  ····················  ····················  ····················
C    4779 hCoV-19/USA/MA-MGH-00460/2020|EPI_ISL_460220|2020-   ····················  ····················  ·····················  ····················  ····················  ····················
C    4780 hCoV-19/USA/MA-MGH-00187/2020|EPI_ISL_460221|2020-   ····················  ····················  ·····················  ····················  ····················  ····················
C    4781 hCoV-19/USA/MA-MGH-00454/2020|EPI_ISL_460223|2020-   ····················  ····················  ·····················  ····················  ····················  ····················
C    4782 hCoV-19/USA/MA-MGH-00521/2020|EPI_ISL_460224|2020-   ····················  ····················  ·····················  ····················  ····················  ····················
C    4783 hCoV-19/USA/MA-MGH-00475/2020|EPI_ISL_460225|2020-   ····················  ····················  ·····················  ····················  ····················  ····················
C    4784 hCoV-19/USA/MA-MGH-00469/2020|EPI_ISL_460226|2020-   ····················  ····················  ·····················  ····················  ····················  ····················
C    4785 hCoV-19/USA/MA-MGH-00480/2020|EPI_ISL_460227|2020-   ····················  ····················  ·····················  ····················  ····················  ····················
C    4786 hCoV-19/USA/MA-MGH-00185/2020|EPI_ISL_460229|2020-   ····················  ····················  ·····················  ····················  ····················  ····················
C    4787 hCoV-19/USA/MA-MGH-00474/2020|EPI_ISL_460230|2020-   ····················  ····················  ·····················  ····················  ····················  ····················
C    4788 hCoV-19/USA/MA-MGH-00680/2020|EPI_ISL_460231|2020-   ····················  ····················  ·····················  ····················  ····················  ····················
C    4789 hCoV-19/USA/MA-MGH-00555/2020|EPI_ISL_460232|2020-   ····················  ····················  ·····················  ····················  ····················  ····················
C    4790 hCoV-19/USA/MA-MGH-00704/2020|EPI_ISL_460233|2020-   ····················  ····················  ·····················  ····················  ····················  ····················
```

```
C  4791 hCoV-19/USA/MA-MGH-00287/2020|EPI_ISL_460234|2020-  ···················· ···················· ···················· ···················· ···················· ····················
C  4792 hCoV-19/USA/MA-MGH-00431/2020|EPI_ISL_460235|2020-  ···················· ···················· ···················· ···················· ···················· ····················
C  4793 hCoV-19/USA/MA-MGH-00239/2020|EPI_ISL_460237|2020-  ···················· ···················· ···················· ···················· ···················· ····················
C  4794 hCoV-19/USA/MA-MGH-00151/2020|EPI_ISL_460238|2020-  ···················· ···················· ···················· ···················· ···················· ····················
C  4795 hCoV-19/USA/MA-MGH-00437/2020|EPI_ISL_460239|2020-  ···················· ···················· ···················· ···················· ···················· ····················
C  4796 hCoV-19/USA/MA-MGH-00037/2020|EPI_ISL_460240|2020-  ···················· ···················· ···················· ···················· ···················· ····················
C  4797 hCoV-19/USA/MA-MGH-00415/2020|EPI_ISL_460241|2020-  ···················· ···················· ···················· ···················· ···················· ····················
C  4798 hCoV-19/USA/MA-MGH-00206/2020|EPI_ISL_460242|2020-  ···················· ···················· ···················· ···················· ···················· ····················
C  4799 hCoV-19/USA/MA-MGH-00270/2020|EPI_ISL_460243|2020-  ···················· ···················· ···················· ···················· ···················· ····················
C  4800 hCoV-19/USA/MA-MGH-00463/2020|EPI_ISL_460244|2020-  ···················· ···················· ···················· ···················· ···················· ····················
C  4801 hCoV-19/USA/MA-MGH-00420/2020|EPI_ISL_460245|2020-  ···················· ···················· ···················· ···················· ···················· ····················
C  4802 hCoV-19/USA/MA-MGH-00717/2020|EPI_ISL_460246|2020-  ···················· ···················· ···················· ···················· ···················· ····················
C  4803 hCoV-19/USA/MA-MGH-00660/2020|EPI_ISL_460248|2020-  ···················· ···················· ···················· ···················· ···················· ····················
C  4804 hCoV-19/USA/MA-MGH-00273/2020|EPI_ISL_460249|2020-  ···················· ···················· ···················· ···················· ···················· ····················
C  4805 hCoV-19/USA/MA-MGH-00438/2020|EPI_ISL_460250|2020-  ···················· ···················· ···················· ···················· ···················· ····················
C  4806 hCoV-19/USA/MA-MGH-00435/2020|EPI_ISL_460252|2020-  ···················· ···················· ···················· ···················· ···················· ····················
C  4807 hCoV-19/USA/MA-MGH-00064/2020|EPI_ISL_460297|2020-  ···················· ···················· ···················· ···················· ···················· ····················
C  4808 hCoV-19/USA/MA-MGH-00649/2020|EPI_ISL_460298|2020-  ···················· ···················· ···················· ···················· ···················· ····················
C  4809 hCoV-19/USA/MA-MGH-00230/2020|EPI_ISL_460299|2020-  ···················· ···················· ···················· ···················· ···················· ····················
C  4810 hCoV-19/India/GJ-GBRC165/2020|EPI_ISL_461500|2020-  ···················· A··················· ···················· ···················· ···················· ··A·················
C  4811 hCoV-19/India/GJ-GBRC166a/2020|EPI_ISL_461501|2020  ···················· A··················· ···················· ···················· ···················· ··A·················
C  4812 hCoV-19/India/GJ-GBRC166b/2020|EPI_ISL_461502|2020  ···················· A··················· ···················· ···················· ···················· ··A·················
C  4813 hCoV-19/India/GJ-GBRC167a/2020|EPI_ISL_461503|2020  ···················· A··················· ···················· ···················· ···················· ··A·················
C  4814 hCoV-19/India/GJ-GBRC167b/2020|EPI_ISL_461504|2020  ···················· ···················· ···················· ···················· ···················· ····················
C  4815 hCoV-19/India/GJ-GBRC168a/2020|EPI_ISL_461505|2020  ···················· ···A················ ···················· ···················· ···················· ····················
C  4816 hCoV-19/India/GJ-GBRC168b/2020|EPI_ISL_461506|2020  ···················· A··················· ···················· ···················· ···················· ····················
C  4820 hCoV-19/England/BIRM-5F521/2020|EPI_ISL_461513|202  ···················· ···················A ···················· ···················· ···················· ····················  1
C  4824 hCoV-19/England/BIRM-5F5B8/2020|EPI_ISL_461518|202  ···················· ···················· ···················· ···················· ···················· ····················
C  4827 hCoV-19/England/BIRM-5F688/2020|EPI_ISL_461523|202  ···················· ···················· ···················· ···················· ···················· ····················
C  4832 hCoV-19/England/BIRM-5FE44/2020|EPI_ISL_461528|202  ···················· ···················· ···················· ···················· ···················· ····················
C  4833 hCoV-19/England/BIRM-5FE53/2020|EPI_ISL_461529|202  ···················· ···················· ···················· ···········T········ ···················· ····················
C  4836 hCoV-19/England/BIRM-5FEBD/2020|EPI_ISL_461532|202  ···················· ···················· ···················· ···················· ···················· ····················
C  4838 hCoV-19/England/BIRM-5FEEA/2020|EPI_ISL_461535|202  ···················· ···················· ···················· ···················· ···················· ····················
C  4839 hCoV-19/England/BIRM-5FF14/2020|EPI_ISL_461537|202  ···················· ···················· ···················· ···················· ···················· ····················
C  4840 hCoV-19/England/BIRM-5FF41/2020|EPI_ISL_461538|202  ···················· ···················· ···················· ···················· ···················· ····················
C  4843 hCoV-19/England/BIRM-5FF9C/2020|EPI_ISL_461543|202  ···················· ···················· ···················· ···················· ···················· ····················
C  4846 hCoV-19/England/CAMB-1B2E5D/2020|EPI_ISL_461546|20  ···················· ···················· ···················· ···················· ···················· ····················
C  4850 hCoV-19/England/CAMB-1B319F/2020|EPI_ISL_461553|20  ···················· ···················· ···················· ···················· ···················· ····················
C  4851 hCoV-19/England/CAMB-1B3557/2020|EPI_ISL_461561|20  ···················· ···················· ···················· ···················· ···················· ····················
C  4852 hCoV-19/England/CAMB-1B3575/2020|EPI_ISL_461562|20  ···················· ···················· ···················· ···················· ···················· ····················
C  4853 hCoV-19/England/CAMB-1B3593/2020|EPI_ISL_461563|20  ···················· ···················· ···················· ···················· ···················· ····················
C  4854 hCoV-19/England/CAMB-1B3A76/2020|EPI_ISL_461571|20  ···················· ···················· ···················· ···················· ···················· ····················
C  4855 hCoV-19/England/CAMB-1B3A85/2020|EPI_ISL_461572|20  ···················· ···················· ···················· ···················· ···················· ····················
C  4856 hCoV-19/England/CAMB-1B3A94/2020|EPI_ISL_461573|20  ···················· ···················· ···················· ···················· ···················· ····················
C  4857 hCoV-19/England/CAMB-1B3AA3/2020|EPI_ISL_461574|20  ···················· ···················· ···················· ···················· ···················· ····················
C  4858 hCoV-19/England/CAMB-1B3AB2/2020|EPI_ISL_461575|20  ···················· ···················· ···················· ···················· ···················· ····················
C  4859 hCoV-19/England/CAMB-1B3B28/2020|EPI_ISL_461578|20  ···················· ···················· ···················· ···················· ···················· ····················
C  4860 hCoV-19/England/CAMB-1B3B64/2020|EPI_ISL_461579|20  ···················· ···················· ···················· ···················· ···················· ····················
C  4861 hCoV-19/England/CAMB-1B3B73/2020|EPI_ISL_461580|20  ···················· ···················· ···················· ···················· ···················· ····················
C  4862 hCoV-19/England/CAMB-1B3B82/2020|EPI_ISL_461581|20  ···················· ···················· ···················· ···················· ···················· ····················
C  4863 hCoV-19/England/CAMB-1B3C61/2020|EPI_ISL_461582|20  ···················· ···················· ···················· ···················· ···················· ····················
C  4864 hCoV-19/England/CAMB-1B3CE9/2020|EPI_ISL_461583|20  ···················· ···················· ···················· ···················· ···················· ····················
C  4865 hCoV-19/Colombia/CUN-GVI-97718/2020|EPI_ISL_447800  ···················· ···················· ···················· ···················· ···················· ····················
C  4866 hCoV-19/Colombia/MET-GVI-97769/2020|EPI_ISL_447802  ···················· ···················· ···················· ···················· ···················· ····················
C  4867 hCoV-19/Colombia/MET-GVI-97782/2020|EPI_ISL_447803  ···················· ···················· ···················· ···················· ···················· ····················
C  4868 hCoV-19/Colombia/MET-GVI-97783/2020|EPI_ISL_447804  ···················· ···················· ···················· ···················· ···················· ····················
C  4869 hCoV-19/Colombia/BOY-GVI-97795/2020|EPI_ISL_447805  ···················· ···················· ···················· ···················· ···················· ····················
C  4870 hCoV-19/Colombia/HUI-GVI-97855/2020|EPI_ISL_447806  ···················· ···················· ···················· ···················· ···················· ····················
C  4871 hCoV-19/Colombia/HUI-GVI-97875/2020|EPI_ISL_447807  ···················· ···················· ···················· ···················· ···················· ····················
C  4872 hCoV-19/Colombia/TOL-GVI-97921/2020|EPI_ISL_447808  ···················· ···················· ···················· ···················· ···················· ····················
C  4873 hCoV-19/Colombia/RIS-GVI-98006/2020|EPI_ISL_447809  ···················· ···················· ···················· ···················· ···················· ····················
C  4874 hCoV-19/Colombia/RIS-GVI-98011/2020|EPI_ISL_447810  ···················· ···················· ···················· ···················· ···················· ····················
C  4875 hCoV-19/Norway/2200/2020|EPI_ISL_447837|2020-03-20  ···················· ···················· ···················· ···················· ···················· ····················
C  4876 hCoV-19/USA/DC-CDC-0019/2020|EPI_ISL_447840|2020-0  ···················· ···················· ···················· ···················· ···················· ····················
C  4877 hCoV-19/USA/FL-CDC-7619/2020|EPI_ISL_447841|2020-0  ···················· ···················· ···················· ···················· ···················· ····················
C  4878 hCoV-19/USA/IA-CDC-8200/2020|EPI_ISL_447842|2020-0  ···················· ···················· ···················· ···················· ···················· ····················
C  4879 hCoV-19/USA/MD-CDC-0025/2020|EPI_ISL_447843|2020-0  ···················· ···················· ···················· ···················· ···················· ····················
C  4880 hCoV-19/USA/PA-CDC-2908/2020|EPI_ISL_447844|2020-0  ···················· ···················· ···················· ···················· ···················· ····················
C  4881 hCoV-19/England/OXON-AC9B0/2020|EPI_ISL_448500|202  ···················· ···················· ···················· ···················· ···················· ····················
C  4882 hCoV-19/England/OXON-AC9ED/2020|EPI_ISL_448501|202  ···················· ···················· ···················· ···················· ···················· ····················
C  4883 hCoV-19/England/OXON-ACA08/2020|EPI_ISL_448502|202  ···················· ···················· ···················· ···················· ···················· ····················
C  4884 hCoV-19/England/OXON-ACA44/2020|EPI_ISL_448503|202  ···················· ···················· ···················· ···················· ···················· ····················
C  4885 hCoV-19/England/OXON-ACA53/2020|EPI_ISL_448504|202  ···················· ···················· ···················· ···················· ···················· ····················
C  4886 hCoV-19/England/OXON-ACA62/2020|EPI_ISL_448505|202  ···················· ···················· ···················· ···················· ···················· ····················
C  4887 hCoV-19/England/OXON-ACA80/2020|EPI_ISL_448506|202  ···················· ···················· ···················· ···················· ···················· ····················
C  4888 hCoV-19/England/OXON-ACADB/2020|EPI_ISL_448507|202  ···················· ···················· ···················· ···················· ···················· ····················
C  4889 hCoV-19/England/OXON-ACAEA/2020|EPI_ISL_448508|202  ···················· ···················· ···················· ···················· ···················· ····················
C  4890 hCoV-19/England/OXON-ACB41/2020|EPI_ISL_448510|202  ···················· ···················· ···················· ···················· ···················· ····················
```

| | | | | | | | | |
|---|---|---|---|---|---|---|---|---|
| C | 4891 hCoV-19/England/OXON-ACB50/2020\|EPI_ISL_448511\|202 | .................... | .................... | .................... | .................... | .................... | .................... | |
| C | 4892 hCoV-19/England/OXON-ACB6F/2020\|EPI_ISL_448512\|202 | .................... | .............A...... | .................... | .................... | .................... | .................... | |
| C | 4893 hCoV-19/England/OXON-ACB8D/2020\|EPI_ISL_448513\|202 | .................... | ...................A | .................... | .................... | .................... | .................... | 1 |
| C | 4894 hCoV-19/England/OXON-ACB9C/2020\|EPI_ISL_448514\|202 | .................... | .................... | .................... | .................... | .................... | .................... | |
| C | 4895 hCoV-19/England/OXON-ACBAB/2020\|EPI_ISL_448515\|202 | .................... | .................... | .................... | .................... | .................... | .................... | |
| C | 4896 hCoV-19/England/OXON-ACBBA/2020\|EPI_ISL_448516\|202 | .................... | .................... | .................... | .................... | .................... | .................... | |
| C | 4897 hCoV-19/England/OXON-ACBE7/2020\|EPI_ISL_448517\|202 | .................... | .................... | .................... | .................... | .................... | .................... | |
| C | 4898 hCoV-19/England/OXON-ACC02/2020\|EPI_ISL_448518\|202 | .................... | .................... | .................... | .................... | .................... | .................... | |
| C | 4899 hCoV-19/England/OXON-ACC11/2020\|EPI_ISL_448519\|202 | .................... | .................... | .................... | .................... | .................... | .................... | |
| C | 4900 hCoV-19/England/OXON-ACC4E/2020\|EPI_ISL_448520\|202 | .................... | .................... | .................... | .................... | .................... | .................... | |
| C | 4901 hCoV-19/England/OXON-ACC5D/2020\|EPI_ISL_448521\|202 | .................... | .................... | .................... | .................... | .................... | .................... | |
| C | 4902 hCoV-19/England/OXON-ACC6C/2020\|EPI_ISL_448522\|202 | .................... | .................... | .................... | .................... | .................... | .................... | |
| C | 4903 hCoV-19/England/OXON-ACCB7/2020\|EPI_ISL_448525\|202 | .................... | .................... | .................... | .................... | .................... | .................... | |
| C | 4904 hCoV-19/England/OXON-ACCC6/2020\|EPI_ISL_448526\|202 | .................... | .................... | .................... | .................... | .................... | .................... | |
| C | 4905 hCoV-19/England/OXON-ACCD5/2020\|EPI_ISL_448527\|202 | .................... | .................... | .................... | .................... | .................... | .................... | |
| C | 4906 hCoV-19/England/OXON-ACD0F/2020\|EPI_ISL_448529\|202 | .................... | .................... | .................... | .................... | .................... | .................... | |
| C | 4907 hCoV-19/England/OXON-ACD1E/2020\|EPI_ISL_448530\|202 | .................... | .................... | .................... | .................... | .................... | .................... | |
| C | 4908 hCoV-19/England/OXON-ACD2D/2020\|EPI_ISL_448531\|202 | .................... | .................... | .................... | .................... | .................... | .................... | |
| C | 4909 hCoV-19/England/OXON-ACD4B/2020\|EPI_ISL_448532\|202 | .................... | .................... | .................... | .................... | .................... | .................... | |
| C | 4910 hCoV-19/England/OXON-ACD5A/2020\|EPI_ISL_448533\|202 | .................... | .................... | .................... | .................... | .................... | .................... | |
| C | 4911 hCoV-19/England/OXON-ACD69/2020\|EPI_ISL_448534\|202 | .................... | .................... | .................... | .................... | .................... | .................... | |
| C | 4912 hCoV-19/England/OXON-ACD87/2020\|EPI_ISL_448535\|202 | .................... | .................... | .................... | .................... | .................... | .................... | |
| C | 4913 hCoV-19/England/OXON-ACD96/2020\|EPI_ISL_448536\|202 | .................... | .................... | .................... | .................... | .................... | .................... | |
| C | 4914 hCoV-19/England/OXON-ACDA5/2020\|EPI_ISL_448537\|202 | .................... | .................... | .................... | .................... | .................... | .................... | |
| C | 4915 hCoV-19/USA/UT-00301/2020\|EPI_ISL_449801\|2020-03-2 | .................... | .................... | .................... | .................... | .................... | .................... | |
| C | 4916 hCoV-19/USA/UT-00303/2020\|EPI_ISL_449802\|2020-03-2 | .................... | .................... | .................... | .................... | .................... | .................... | |
| C | 4917 hCoV-19/USA/UT-00346/2020\|EPI_ISL_449804\|2020-03-2 | .................... | .................... | .................... | .................... | .................... | .................... | |
| C | 4918 hCoV-19/USA/UT-00352/2020\|EPI_ISL_449805\|2020-03-2 | .................... | .................... | .................... | .................... | .................... | .................... | |
| C | 4919 hCoV-19/USA/UT-00349/2020\|EPI_ISL_449806\|2020-03-2 | .................... | .................... | .................... | .................... | .................... | .................... | |
| C | 4920 hCoV-19/USA/UT-00348/2020\|EPI_ISL_449809\|2020-03-2 | .................... | .................... | .................... | .................... | .................... | .................... | |
| C | 4921 hCoV-19/USA/UT-01560/2020\|EPI_ISL_449815\|2020-04-0 | .................... | .................... | .................... | .................... | .................... | .................... | |
| C | 4922 hCoV-19/USA/UT-01562/2020\|EPI_ISL_449816\|2020-04-0 | .................... | .................... | .................... | .................... | .................... | .................... | |
| C | 4923 hCoV-19/USA/UT-01564/2020\|EPI_ISL_449817\|2020-04-0 | .................... | .................... | .................... | .................... | .................... | .................... | |
| C | 4924 hCoV-19/USA/UT-01566/2020\|EPI_ISL_449818\|2020-04-0 | .................... | .................... | .................... | .................... | .................... | .................... | |
| C | 4925 hCoV-19/USA/UT-01568/2020\|EPI_ISL_449819\|2020-04-0 | .................... | .................... | .................... | .................... | .................... | .................... | |
| C | 4926 hCoV-19/USA/UT-01570/2020\|EPI_ISL_449820\|2020-04-0 | .................... | .................... | .................... | .................... | .................... | .................... | |
| C | 4927 hCoV-19/USA/UT-01572/2020\|EPI_ISL_449821\|2020-04-0 | .................... | .................... | .................... | .................... | .................... | .................... | |
| C | 4928 hCoV-19/USA/UT-01573/2020\|EPI_ISL_449822\|2020-04-0 | .................... | .................... | .................... | .................... | .................... | .................... | |
| C | 4929 hCoV-19/USA/UT-01575/2020\|EPI_ISL_449823\|2020-04-0 | .................... | .................... | .................... | .................... | .................... | .................... | |
| C | 4930 hCoV-19/USA/UT-01578/2020\|EPI_ISL_449824\|2020-04-0 | .................... | .................... | .................... | .................... | .................... | .................... | |
| C | 4931 hCoV-19/USA/UT-01579/2020\|EPI_ISL_449825\|2020-04-0 | .................... | .................... | .................... | .................... | .................... | .................... | |
| C | 4932 hCoV-19/USA/UT-01581/2020\|EPI_ISL_449826\|2020-04-0 | .................... | .................... | .................... | .................... | .................... | .................... | |
| C | 4933 hCoV-19/USA/UT-01582/2020\|EPI_ISL_449827\|2020-04-0 | .................... | .................... | .................... | .................... | .................... | .................... | |
| C | 4934 hCoV-19/USA/UT-01583/2020\|EPI_ISL_449828\|2020-04-0 | .................... | .................... | .................... | .................... | .................... | .................... | |
| C | 4935 hCoV-19/USA/UT-01585/2020\|EPI_ISL_449829\|2020-04-0 | .................... | .................... | .................... | .................... | .................... | .................... | |
| C | 4936 hCoV-19/USA/UT-01587/2020\|EPI_ISL_449830\|2020-04-0 | .................... | .................... | .................... | .................... | .................... | .................... | |
| C | 4937 hCoV-19/USA/UT-01590/2020\|EPI_ISL_449831\|2020-04-0 | .................... | .................... | .................... | .................... | .................... | .................... | |
| C | 4938 hCoV-19/USA/UT-01595/2020\|EPI_ISL_449833\|2020-04-0 | .................... | .................... | .................... | .................... | .................... | .................... | |
| C | 4939 hCoV-19/USA/UT-01598/2020\|EPI_ISL_449834\|2020-04-0 | .................... | .................... | .................... | .................... | .................... | .................... | |
| C | 4940 hCoV-19/USA/UT-01601/2020\|EPI_ISL_449835\|2020-04-0 | .................... | .................... | .................... | .................... | .................... | .................... | |
| C | 4941 hCoV-19/USA/UT-01602/2020\|EPI_ISL_449836\|2020-04-0 | .................... | .................... | .................... | .................... | .................... | .................... | |
| C | 4942 hCoV-19/USA/UT-01607/2020\|EPI_ISL_449837\|2020-04-0 | .................... | .................... | .................... | .................... | .................... | .................... | |
| C | 4943 hCoV-19/USA/UT-01609/2020\|EPI_ISL_449838\|2020-04-0 | .................... | .................... | .................... | .................... | .................... | .................... | |
| C | 4944 hCoV-19/USA/WA-S669/2020\|EPI_ISL_449839\|2020-04-08 | .................... | .................... | .................... | .................... | .................... | .................... | |
| C | 4945 hCoV-19/USA/WA-S670/2020\|EPI_ISL_449840\|2020-04-08 | .................... | .................... | .................... | .................... | .................... | .................... | |
| C | 4946 hCoV-19/USA/WA-S671/2020\|EPI_ISL_449841\|2020-04-08 | .................... | .................... | .................... | .................... | .................... | .................... | |
| C | 4947 hCoV-19/USA/WA-S672/2020\|EPI_ISL_449842\|2020-04-08 | .................... | .................... | .................... | .................... | .................... | .................... | |
| C | 4948 hCoV-19/USA/WA-S673/2020\|EPI_ISL_449843\|2020-04-08 | .................... | .................... | .................... | .................... | .................... | .................... | |
| C | 4949 hCoV-19/USA/WA-S674/2020\|EPI_ISL_449844\|2020-04-08 | .................... | .................... | .................... | .................... | .................... | .................... | |
| C | 4950 hCoV-19/USA/WA-S675/2020\|EPI_ISL_449845\|2020-04-08 | .................... | .................... | .................... | .................... | .................... | .................... | |
| C | 4951 hCoV-19/USA/WA-S676/2020\|EPI_ISL_449846\|2020-04-07 | .................... | .................... | .................... | .................... | .................... | .................... | |
| C | 4952 hCoV-19/USA/WA-S677/2020\|EPI_ISL_449847\|2020-04-07 | .................... | .................... | .................... | .................... | .................... | .................... | |
| C | 4953 hCoV-19/USA/WA-S678/2020\|EPI_ISL_449848\|2020-04-07 | .................... | .................... | .................... | .................... | .................... | .................... | |
| C | 4954 hCoV-19/USA/WA-S679/2020\|EPI_ISL_449849\|2020-04-07 | .................... | .................... | .................... | .................... | .................... | .................... | |
| C | 4955 hCoV-19/USA/WA-S680/2020\|EPI_ISL_449850\|2020-04-08 | .................... | .................... | .................... | .................... | .................... | .................... | |
| C | 4956 hCoV-19/USA/WA-S681/2020\|EPI_ISL_449851\|2020-04-08 | .................... | .................... | .................... | .................... | .................... | .................... | |
| C | 4957 hCoV-19/USA/WA-S682/2020\|EPI_ISL_449852\|2020-04-09 | .................... | .................... | .................... | .................... | .................... | .................... | |
| C | 4958 hCoV-19/USA/WA-S683/2020\|EPI_ISL_449853\|2020-04-07 | .................... | .................... | .................... | .................... | .................... | .................... | |
| C | 4959 hCoV-19/USA/WA-S684/2020\|EPI_ISL_449854\|2020-04-11 | .................... | .................... | .................... | .................... | .................... | .................... | |
| C | 4960 hCoV-19/USA/WA-S685/2020\|EPI_ISL_449855\|2020-04-11 | .................... | .................... | .................... | .................... | .................... | .................... | |
| C | 4961 hCoV-19/USA/WA-S686/2020\|EPI_ISL_449856\|2020-04-08 | .................... | .................... | .................... | .................... | .................... | .................... | |
| C | 4962 hCoV-19/USA/WA-S687/2020\|EPI_ISL_449857\|2020-04-08 | .................... | .................... | .................... | .................... | .................... | .................... | |
| C | 4963 hCoV-19/USA/WA-S688/2020\|EPI_ISL_449858\|2020-04-08 | .................... | .................... | .................... | .................... | .................... | .................... | |
| C | 4964 hCoV-19/USA/WA-S689/2020\|EPI_ISL_449859\|2020-04-08 | .................... | .................... | .................... | .................... | .................... | .................... | |
| C | 4965 hCoV-19/USA/WA-S690/2020\|EPI_ISL_449860\|2020-04-08 | .................... | .................... | .................... | .................... | .................... | .................... | |
| C | 4966 hCoV-19/USA/WA-S691/2020\|EPI_ISL_449861\|2020-04-08 | .................... | .................... | .................... | .................... | .................... | .................... | |
| C | 4967 hCoV-19/USA/WA-S692/2020\|EPI_ISL_449862\|2020-04-08 | .................... | .................... | .................... | .................... | .................... | .................... | |
| C | 4968 hCoV-19/USA/WA-S693/2020\|EPI_ISL_449863\|2020-04-08 | .................... | .................... | .................... | .................... | .................... | .................... | |

C 4969 hCoV-19/USA/WA-S694/2020|EPI_ISL_449864|2020-04-08 ···················· ···················· ····················· ···················· ···················· ····················
C 4970 hCoV-19/USA/WA-S695/2020|EPI_ISL_449865|2020-04-08 ···················· ···················· ····················· ···················· ···················· ····················
C 4971 hCoV-19/USA/WA-S696/2020|EPI_ISL_449866|2020-04-09 ···················· ···················· ····················· ···················· ···················· ····················
C 4972 hCoV-19/USA/WA-S697/2020|EPI_ISL_449867|2020-04-08 ···················· ···················· ····················· ···················· ···················· ····················
C 4973 hCoV-19/USA/WA-S698/2020|EPI_ISL_449868|2020-04-08 ···················· ···················· ····················· ···················· ···················· ····················
C 4974 hCoV-19/USA/GA-CDC-4518/2020|EPI_ISL_450801|2020-0 ···················· ···················· ····················· ···················· ···················· ····················
C 4975 hCoV-19/USA/PA-CDC-8398/2020|EPI_ISL_450802|2020-0 ···················· ···················· ····················· ···················· ···················· ····················
C 4976 hCoV-19/USA/PA-CDC-9081/2020|EPI_ISL_450803|2020-0 ···················· ···················· ····················· ···················· ···················· ····················
C 4977 hCoV-19/USA/VI-CDC-3810/2020|EPI_ISL_450804|2020-0 ···················· ···················· ····················· ···················· ···················· ····················
C 4978 hCoV-19/USA/VI-CDC-3943/2020|EPI_ISL_450806|2020-0 ···················· ···················· ····················· ···················· ···················· ····················
C 4979 hCoV-19/Sweden/20-06097/2020|EPI_ISL_450807|2020-0 ···················· ···················· ····················· ···················· ···················· ····················
C 4980 hCoV-19/Sweden/20-06665/2020|EPI_ISL_450808|2020-0 ···················· ···················· ····················· ···················· ···················· ····················
C 4981 hCoV-19/Sweden/20-06695/2020|EPI_ISL_450809|2020-0 ···················· ···················· ····················· ···················· ···················· ····················
C 4982 hCoV-19/Sweden/20-06737/2020|EPI_ISL_450810|2020-0 ···················· ···················· ····················· ···················· ···················· ····················
C 4983 hCoV-19/Sweden/20-06909/2020|EPI_ISL_450811|2020-0 ···················· ···················· ····················· ···················· ···················· ····················
C 4984 hCoV-19/Sweden/20-07041/2020|EPI_ISL_450812|2020-0 ···················· ···················· ····················· ···················· ···················· ····················
C 4985 hCoV-19/Sweden/20-07299/2020|EPI_ISL_450813|2020-0 ···················· ···················· ····················· ···················· ···················· ····················
C 4986 hCoV-19/Sweden/20-07734/2020|EPI_ISL_450814|2020-0 ···················· ···················· ····················· ···················· ···················· ····················
C 4987 hCoV-19/Sweden/20-07737/2020|EPI_ISL_450815|2020-0 ···················· ···················· ····················· ···················· ···················· ····················
C 4988 hCoV-19/Sweden/20-07739/2020|EPI_ISL_450816|2020-0 ···················· ···················· ····················· ···················· ···················· ····················
C 4989 hCoV-19/Sweden/20-07743/2020|EPI_ISL_450817|2020-0 ···················· ···················· ····················· ···················· ···················· ····················
C 4990 hCoV-19/Sweden/20-07768/2020|EPI_ISL_450818|2020-0 ···················· ···················· ····················· ···················· ···················· ····················
C 4991 hCoV-19/Sweden/20-07833/2020|EPI_ISL_450819|2020-0 ···················· ···················· ····················· ···················· ···················· ····················
C 4992 hCoV-19/Sweden/20-07847/2020|EPI_ISL_450820|2020-0 ···················· ···················· ····················· ···················· ···················· ····················
C 4993 hCoV-19/Sweden/20-07860/2020|EPI_ISL_450821|2020-0 ···················· ···················· ····················· ···················· ···················· ····················
C 4994 hCoV-19/Sweden/20-07861/2020|EPI_ISL_450822|2020-0 ···················· ···················· ····················· ···················· ···················· ····················
C 4995 hCoV-19/Sweden/20-07919/2020|EPI_ISL_450823|2020-0 ···················· ···················· ····················· ···················· ···················· ····················
C 4996 hCoV-19/Sweden/20-07928/2020|EPI_ISL_450824|2020-0 ···················· ···················· ····················· ···················· ···················· ····················
C 4997 hCoV-19/Sweden/20-14084/2020|EPI_ISL_450825|2020-0 ···················· ···················· ····················· ···················· ···················· ····················
C 4998 hCoV-19/Sweden/20-14087/2020|EPI_ISL_450826|2020-0 ···················· ···················· ····················· ···················· ···················· ····················
C 4999 hCoV-19/Sweden/20-14089/2020|EPI_ISL_450827|2020-0 ···················· ···················· ····················· ···················· ···················· ····················
C 5000 hCoV-19/Sweden/20-14090/2020|EPI_ISL_450828|2020-0 ···················· ···················· ····················· ···················· ···················· ····················
C 5001 hCoV-19/Sweden/20-14094/2020|EPI_ISL_450829|2020-0 ···················· ···················· ····················· ···················· ···················· ····················
C 5002 hCoV-19/Sweden/20-51023/2020|EPI_ISL_450830|2020-0 ···················· ···················· ····················· ···················· ···················· ····················
C 5003 hCoV-19/Sweden/20-51044/2020|EPI_ISL_450831|2020-0 ···················· ···················· ····················· ···················· ···················· ····················
C 5004 hCoV-19/Sweden/20-51167/2020|EPI_ISL_450832|2020-0 ···················· ···················· ····A················ ···················· ···················· ····················
C 5005 hCoV-19/Sweden/20-51311/2020|EPI_ISL_450833|2020-0 ···················· ···················· ····················· ···················· ···················· ····················
C 5006 hCoV-19/Sweden/20-08168/2020|EPI_ISL_450834|2020-0 ···················· ···················· ····················· ···················· ···················· ····················
C 5007 hCoV-19/Sweden/20-08352/2020|EPI_ISL_450835|2020-0 ···················· ···················· ····················· ···················· ···················· ····················
C 5008 hCoV-19/Sweden/20-08715/2020|EPI_ISL_450836|2020-0 ···················· ···················· ····················· ···················· ···················· ····················
C 5009 hCoV-19/Sweden/20-08718/2020|EPI_ISL_450837|2020-0 ···················· ···················· ····················· ···················· ···················· ····················
C 5010 hCoV-19/Bangladesh/DU-50740/2020|EPI_ISL_450839|20 ···················· ···················· ····················· ···················· ···················· ····················
C 5011 hCoV-19/Bangladesh/DU-50758/2020|EPI_ISL_450840|20 ···················· ···················· ····················· ···················· ···················· ····················
C 5012 hCoV-19/Bangladesh/DU-CARS-5586/2020|EPI_ISL_45084 ···················· ···················· ····················· ···················· ···················· ····················
C 5013 hCoV-19/Bangladesh/DU-50768/2020|EPI_ISL_450843|20 ···················· ···················· ····················· ···················· ···················· ····················
C 5014 hCoV-19/USA/FL-BPHL-0115/2020|EPI_ISL_450849|2020- ···················· ···················· ····················· ···················· ···················· ····················
C 5015 hCoV-19/Australia/SAP034/2020|EPI_ISL_451079|2020- ···················· ···················· ····················· ···················· ···················· ····················
C 5016 hCoV-19/Australia/SAP035/2020|EPI_ISL_451080|2020- ···················· ···················· ····················· ···················· ···················· ····················
C 5017 hCoV-19/Australia/SAP037/2020|EPI_ISL_451081|2020- ···················· ···················· ····················· ···················· ···················· ····················
C 5018 hCoV-19/Australia/SAP041/2020|EPI_ISL_451082|2020- ···················· ···················· ····················· ···················· ···················· ····················
C 5019 hCoV-19/Australia/SAP042/2020|EPI_ISL_451083|2020- ···················· ···················· ····················· ···················· ···················· ····················
C 5020 hCoV-19/Australia/SAP044/2020|EPI_ISL_451084|2020- ···················· ···················· ····················· ···················· ···················· ····················
C 5021 hCoV-19/Australia/SAP048/2020|EPI_ISL_451087|2020- ···················· ···················· ····················· ···················· ···················· ····················
C 5022 hCoV-19/Australia/SAP049/2020|EPI_ISL_451088|2020- ···················· ···················· ····················· ···················· ···················· ····················
C 5023 hCoV-19/Australia/SAP050/2020|EPI_ISL_451089|2020- ···················· ···················· ····················· ···················· ···················· ····················
C 5024 hCoV-19/Australia/SAP051/2020|EPI_ISL_451090|2020- ···················· ···················· ····················· ···················· ···················· ····················
C 5025 hCoV-19/Australia/SAP052/2020|EPI_ISL_451091|2020- ···················· ···················· ····················· ···················· ···················· ····················
C 5026 hCoV-19/Australia/SAP054/2020|EPI_ISL_451093|2020- ···················· ···················· ····················· ···················· ···················· ····················
C 5027 hCoV-19/Australia/SAP055/2020|EPI_ISL_451094|2020- ···················· ···················· ····················· ···················· ···················· ····················
C 5028 hCoV-19/Australia/SAP056/2020|EPI_ISL_451095|2020- ···················· ···················· ····················· ···················· ···················· ····················
C 5029 hCoV-19/Australia/SAP057/2020|EPI_ISL_451096|2020- ···················· ···················· ····················· ···················· ···················· ····················
C 5030 hCoV-19/Australia/SAP058/2020|EPI_ISL_451097|2020- ···················· ···················· ····················· ···················· ···················· ····················
C 5031 hCoV-19/Australia/SAP059/2020|EPI_ISL_451098|2020- ···················· ···················· ····················· ···················· ···················· ····················
C 5032 hCoV-19/Australia/SAP060/2020|EPI_ISL_451099|2020- ···················· ···················· ····················· ···················· ···················· ····················
C 5033 hCoV-19/Beijing/DT-travelUK01/2020|EPI_ISL_452355| ···················· ···················· ····················· ···················· ···················· ····················
C 5034 hCoV-19/Beijing/DT-travelUK03/2020|EPI_ISL_452356| ···················· ···················· ····················· ···················· ···················· ····················
C 5035 hCoV-19/Beijing/DT-WH02/2020|EPI_ISL_452357|2020-0 ···················· ···················· ····················· ···················· ···················· ····················
C 5036 hCoV-19/Beijing/DT-BJ01/2020|EPI_ISL_452358|2020-0 ···················· ···················· ····················· ···················· ···················· ····················
C 5037 hCoV-19/Beijing/DT-BJ02/2020|EPI_ISL_452359|2020-0 ···················· ···················· ····················· ···················· ···················· ····················
C 5038 hCoV-19/Beijing/DT-WH01/2020|EPI_ISL_452360|2020-0 ···················· ···················· ····················· ···················· ···················· ····················
C 5039 hCoV-19/Beijing/DT-BJ03/2020|EPI_ISL_452361|2020-0 ···················· ···················· ····················· ···················· ···················· ····················
C 5040 hCoV-19/Beijing/DT-WH03/2020|EPI_ISL_452362|2020-0 ···················· ···················· ····················· ···················· ···················· ····················
C 5041 hCoV-19/Beijing/DT-WH04/2020|EPI_ISL_452363|2020-0 ···················· ···················· ····················· ···················· ···················· ····················
C 5042 hCoV-19/Beijing/DT-BJ04/2020|EPI_ISL_452364|2020-0 ···················· ···················· ····················· ···················· ···················· ····················
C 5044 hCoV-19/Spain/AN-IBV-002058/2020|EPI_ISL_452371|20 ···················· ···················· ····················· ···················· ···················· ····················
C 5045 hCoV-19/Spain/AN-IBV-002057/2020|EPI_ISL_452372|20 ···················· ···················· ····················· ···················· ···················· ····················
C 5059 hCoV-19/Scotland/CVR2491/2020|EPI_ISL_453056|2020- ···················· ···················· ····················· ···················· ···················· ····················
C 5060 hCoV-19/Scotland/CVR2557/2020|EPI_ISL_453060|2020- ···················· ···················· ····················· ···················· ···················· ····················

| | | | | | | | |
|---|---|---|---|---|---|---|---|
| C | 5061 hCoV-19/Scotland/CVR2595/2020\|EPI_ISL_453061\|2020- | ···················· | ···················· | ···················· | ···················· | ···················· | ···················· |
| C | 5062 hCoV-19/Scotland/CVR2596/2020\|EPI_ISL_453062\|2020- | ···················· | ···················· | ···················· | ···················· | ···················· | ···················· |
| C | 5063 hCoV-19/Scotland/CVR2601/2020\|EPI_ISL_453063\|2020- | ···················· | ···················· | ···················· | ···················· | ···················· | ···················· |
| C | 5064 hCoV-19/Scotland/CVR2641/2020\|EPI_ISL_453065\|2020- | ···················· | ···················· | ···················· | ···················· | ···················· | ···················· |
| C | 5065 hCoV-19/Scotland/CVR2676/2020\|EPI_ISL_453069\|2020- | ···················· | ···················· | ···················· | ···················· | ···················· | ···················· |
| C | 5066 hCoV-19/Scotland/CVR2687/2020\|EPI_ISL_453070\|2020- | ···················· | ···················· | ···················· | ···················· | ···················· | ···················· |
| C | 5067 hCoV-19/Scotland/CVR2732/2020\|EPI_ISL_453072\|2020- | ···················· | ···················· | ···················· | ···················· | ···················· | ···················· |
| C | 5068 hCoV-19/Scotland/CVR2735/2020\|EPI_ISL_453074\|2020- | ···················· | ···················· | ···················· | ···················· | ···················· | ···················· |
| C | 5069 hCoV-19/Scotland/CVR2794/2020\|EPI_ISL_453075\|2020- | ···················· | ···················· | ···················· | ···················· | ···················· | ···················· |
| C | 5070 hCoV-19/Scotland/CVR2795/2020\|EPI_ISL_453076\|2020- | ···················· | ···················· | ··········T········· | ···················· | ···················· | ···················· |
| C | 5071 hCoV-19/Scotland/CVR2798/2020\|EPI_ISL_453078\|2020- | ···················· | ···················· | ···················· | ···················· | ···················· | ···················· |
| C | 5073 hCoV-19/Scotland/CVR2889/2020\|EPI_ISL_453083\|2020- | ···················· | ·A·················· | ···················· | ···················· | ···················· | ···················· |
| C | 5074 hCoV-19/Scotland/CVR2890/2020\|EPI_ISL_453084\|2020- | ···················· | ·A·················· | ···················· | ···················· | ···················· | ···················· |
| C | 5075 hCoV-19/Scotland/CVR2892/2020\|EPI_ISL_453085\|2020- | ···················· | ·A·················· | ···················· | ···················· | ···················· | ···················· |
| C | 5076 hCoV-19/Scotland/CVR2967/2020\|EPI_ISL_453086\|2020- | ···················· | ···················· | ···················· | ···················· | ···················· | ···················· |
| C | 5077 hCoV-19/Scotland/CVR2969/2020\|EPI_ISL_453088\|2020- | ···················· | ···················· | ···················· | ···················· | ···················· | ···················· |
| C | 5078 hCoV-19/Scotland/CVR3005/2020\|EPI_ISL_453090\|2020- | ···················· | ···················· | ···················· | ···················· | ···················· | ···················· |
| C | 5080 hCoV-19/Scotland/EDB2593/2020\|EPI_ISL_453099\|2020- | ···················· | ·A·················· | ···················· | ···················· | ···················· | ···················· |
| C | 5081 hCoV-19/England/PORT-2CF4D2/2020\|EPI_ISL_453687\|20 | ···················· | ···················· | ···················· | ···················· | ···················· | ···················· |
| C | 5082 hCoV-19/England/PORT-2CF52A/2020\|EPI_ISL_453691\|20 | ···················· | ···················· | ···················· | ···················· | ···················· | ···················· |
| C | 5083 hCoV-19/England/SHEF-C0A95/2020\|EPI_ISL_453696\|202 | ···················· | ···················· | ···················· | ···················· | ···················· | ···················· |
| C | 5084 hCoV-19/England/SHEF-C1C16/2020\|EPI_ISL_453698\|202 | ···················· | ···················· | ···················· | ···················· | ···················· | ···················· |
| C | 5085 hCoV-19/England/SHEF-C225F/2020\|EPI_ISL_453699\|202 | ···················· | ···················· | ···················· | ···················· | ···················· | ···················· |
| C | 5086 hCoV-19/USA/PA-MGEL99/2020\|EPI_ISL_454379\|2020-04- | ···················· | ···················· | ···················· | ···················· | ···················· | ···················· |
| C | 5087 hCoV-19/USA/PA-MGEL100/2020\|EPI_ISL_454380\|2020-04 | ···················· | ···················· | ···················· | ···················· | ···················· | ···················· |
| C | 5088 hCoV-19/USA/PA-MGEL102/2020\|EPI_ISL_454381\|2020-04 | ···················· | ···················· | ···················· | ···················· | ···················· | ···················· |
| C | 5089 hCoV-19/USA/PA-MGEL103/2020\|EPI_ISL_454382\|2020-03 | ···················· | ···················· | ···················· | ···················· | ···················· | ···················· |
| C | 5090 hCoV-19/USA/PA-MGEL104/2020\|EPI_ISL_454383\|2020-03 | ···················· | ···················· | ···················· | ···················· | ···················· | ···················· |
| C | 5091 hCoV-19/USA/PA-MGEL107/2020\|EPI_ISL_454384\|2020-03 | ···················· | ···················· | ···················· | ···················· | ···················· | ···················· |
| C | 5092 hCoV-19/USA/PA-MGEL108/2020\|EPI_ISL_454385\|2020 | ···················· | ···················· | ···················· | ···················· | ···················· | ···················· |
| C | 5093 hCoV-19/USA/PA-MGEL109/2020\|EPI_ISL_454386\|2020 | ···················· | ···················· | ···················· | ···················· | ···················· | ···················· |
| C | 5094 hCoV-19/USA/PA-MGEL110/2020\|EPI_ISL_454387\|2020-04 | ···················· | ···················· | ···················· | ···················· | ···················· | ···················· |
| C | 5095 hCoV-19/USA/PA-MGEL111/2020\|EPI_ISL_454388\|2020 | ···················· | ···················· | ···················· | ···················· | ···················· | ···················· |
| C | 5096 hCoV-19/USA/PA-MGEL113/2020\|EPI_ISL_454389\|2020 | ···················· | ···················· | ···················· | ···················· | ···················· | ···················· |
| C | 5097 hCoV-19/USA/PA-MGEL114/2020\|EPI_ISL_454390\|2020 | ···················· | ···················· | ···················· | ···················· | ···················· | ···················· |
| C | 5098 hCoV-19/USA/PA-MGEL115/2020\|EPI_ISL_454391\|2020 | ···················· | ···················· | ···················· | ···················· | ···················· | ···················· |
| C | 5099 hCoV-19/USA/PA-MGEL118/2020\|EPI_ISL_454392\|2020 | ···················· | ···················· | ···················· | ···················· | ···················· | ···················· |
| C | 5100 hCoV-19/USA/PA-MGEL119/2020\|EPI_ISL_454393\|2020 | ···················· | ···················· | ···················· | ···················· | ···················· | ···················· |
| C | 5101 hCoV-19/USA/PA-MGEL122/2020\|EPI_ISL_454394\|2020 | ···················· | ···················· | ···················· | ···················· | ···················· | ···················· |
| C | 5102 hCoV-19/USA/PA-MGEL123/2020\|EPI_ISL_454395\|2020 | ···················· | ···················· | ···················· | ···················· | ···················· | ···················· |
| C | 5103 hCoV-19/USA/PA-MGEL125/2020\|EPI_ISL_454396\|2020 | ···················· | ···················· | ···················· | ···················· | ···················· | ···················· |
| C | 5104 hCoV-19/USA/PA-MGEL128/2020\|EPI_ISL_454397\|2020 | ···················· | ···················· | ···················· | ···················· | ···················· | ···················· |
| C | 5105 hCoV-19/USA/PA-MGEL130/2020\|EPI_ISL_454398\|2020 | ···················· | ···················· | ···················· | ···················· | ···················· | ···················· |
| C | 5106 hCoV-19/USA/PA-MGEL134/2020\|EPI_ISL_454399\|2020-04 | ···················· | ···················· | ···················· | ···················· | ···················· | ···················· |
| C | 5107 hCoV-19/Netherlands/Overijssel_11/2020\|EPI_ISL_460 | ···················· | ···················· | ···················· | ···················· | ···················· | ···················· |
| C | 5108 hCoV-19/Netherlands/Overijssel_12/2020\|EPI_ISL_460 | ···················· | ···················· | ···················· | ···················· | ···················· | ···················· |
| C | 5109 hCoV-19/Netherlands/Overijssel_13/2020\|EPI_ISL_460 | ···················· | ···················· | ···················· | ···················· | ···················· | ···················· |
| C | 5110 hCoV-19/Netherlands/Overijssel_14/2020\|EPI_ISL_460 | ···················· | ···················· | ···················· | ···················· | ···················· | ···················· |
| C | 5111 hCoV-19/Netherlands/Overijssel_15/2020\|EPI_ISL_460 | ···················· | ···················· | ···················· | ···················· | ···················· | ···················· |
| C | 5112 hCoV-19/Netherlands/Overijssel_16/2020\|EPI_ISL_460 | ···················· | ···················· | ···················· | ···················· | ···················· | ···················· |
| C | 5113 hCoV-19/Netherlands/Overijssel_17/2020\|EPI_ISL_460 | ···················· | ···················· | ···················· | ···················· | ···················· | ···················· |
| C | 5114 hCoV-19/Netherlands/Overijssel_18/2020\|EPI_ISL_460 | ···················· | ···················· | ···················· | ···················· | ···················· | ···················· |
| C | 5115 hCoV-19/Netherlands/Overijssel_19/2020\|EPI_ISL_460 | ···················· | ···················· | ···················· | ···················· | ···················· | ···················· |
| C | 5116 hCoV-19/Netherlands/Overijssel_20/2020\|EPI_ISL_460 | ···················· | ···················· | ···················· | ···················· | ···················· | ···················· |
| C | 5117 hCoV-19/Tunisia/COV1339/2020\|EPI_ISL_463003\|2020-0 | ···················· | ···················· | ···················· | ···················· | ···················· | ············G······· |
| C | 5118 hCoV-19/Tunisia/COV1663/2020\|EPI_ISL_463004\|2020-0 | ···················· | ···················· | ···················· | ···················· | ···················· | ···················· |
| C | 5119 hCoV-19/Tunisia/COV0425/2020\|EPI_ISL_463005\|2020-0 | ···················· | ···················· | ···················· | ···················· | ···················· | ···················· |
| C | 5120 hCoV-19/Tunisia/COV1482/2020\|EPI_ISL_463006\|2020-0 | ···················· | ···················· | ···················· | ···················· | ···················· | ···················· |
| C | 5121 hCoV-19/Colombia/CUN-GVI-98087/2020\|EPI_ISL_447812 | ···················· | ···················· | ···················· | ···················· | ···················· | ···················· |
| C | 5122 hCoV-19/Colombia/CUN-GVI-98290/2020\|EPI_ISL_447813 | ···················· | ···················· | ···················· | ···················· | ···················· | ···················· |
| C | 5123 hCoV-19/Colombia/COR-GVI-97517/2020\|EPI_ISL_447816 | ···················· | ·A·················· | ···················· | ···················· | ···················· | ···················· |
| C | 5124 hCoV-19/Colombia/COR-GVI-97527/2020\|EPI_ISL_447817 | ···················· | ···················· | ···················· | ···················· | ···················· | ···················· |
| C | 5125 hCoV-19/Sichuan/SC-WCH2-006/2020\|EPI_ISL_451076\|20 | ···················· | ···················· | ···················· | ···················· | ···················· | ···················· |
| C | 5126 hCoV-19/Australia/SAP032/2020\|EPI_ISL_451077\|2020- | ···················· | ···················· | ···················· | ···················· | ···················· | ···················· |
| C | 5127 hCoV-19/Australia/SAP033/2020\|EPI_ISL_451078\|2020- | ···················· | ···················· | ···················· | ···················· | ···················· | ···················· |
| C | 5128 hCoV-19/USA/MI-MDHHS-SC20426/2020\|EPI_ISL_452300\|2 | ···················· | ···················· | ···················· | ···················· | ···················· | ···················· |
| C | 5129 hCoV-19/USA/MI-MDHHS-SC20427/2020\|EPI_ISL_452301\|2 | ···················· | ···················· | ···················· | ···················· | ···················· | ···················· |
| C | 5130 hCoV-19/USA/MI-MDHHS-SC20428/2020\|EPI_ISL_452302\|2 | ···················· | ···················· | ···················· | ···················· | ···················· | ···················· |
| C | 5131 hCoV-19/USA/MI-MDHHS-SC20429/2020\|EPI_ISL_452303\|2 | ···················· | ···················· | ···················· | ···················· | ···················· | ···················· |
| C | 5132 hCoV-19/USA/MI-MDHHS-SC20431/2020\|EPI_ISL_452305\|2 | ···················· | ···················· | ···················· | ···················· | ···················· | ···················· |
| C | 5133 hCoV-19/USA/MI-MDHHS-SC20432/2020\|EPI_ISL_452306\|2 | ···················· | ···················· | ···················· | ···················· | ···················· | ···················· |
| C | 5134 hCoV-19/USA/MI-MDHHS-SC20435/2020\|EPI_ISL_452309\|2 | ···················· | ···················· | ···················· | ···················· | ···················· | ···················· |
| C | 5135 hCoV-19/USA/MI-MDHHS-SC20436/2020\|EPI_ISL_452310\|2 | ···················· | ···················· | ···················· | ···················· | ···················· | ···················· |
| C | 5136 hCoV-19/USA/MI-MDHHS-SC20437/2020\|EPI_ISL_452311\|2 | ···················· | ···················· | ···················· | ···················· | ···················· | ···················· |
| C | 5137 hCoV-19/USA/MI-MDHHS-SC20442/2020\|EPI_ISL_452316\|2 | ···················· | ···················· | ···················· | ···················· | ···················· | ···················· |
| C | 5138 hCoV-19/USA/MI-MDHHS-SC20443/2020\|EPI_ISL_452317\|2 | ···················· | ···················· | ···················· | ···················· | ···················· | ···················· |
| C | 5139 hCoV-19/USA/MI-MDHHS-SC20444/2020\|EPI_ISL_452318\|2 | ···················· | ···················· | ···················· | ···················· | ···················· | ···················· |
| C | 5140 hCoV-19/USA/MI-MDHHS-SC20445/2020\|EPI_ISL_452319\|2 | ···················· | ···················· | ···················· | ···················· | ···················· | ···················· |

| | | | | | | | | |
|---|---|---|---|---|---|---|---|---|
| C | 5141 | hCoV-19/USA/MI-MDHHS-SC20446/2020\|EPI_ISL_452320\|2 | .................... | .................... | .................... | .................... | .................... | .................... |
| C | 5142 | hCoV-19/USA/MI-MDHHS-SC20447/2020\|EPI_ISL_452321\|2 | .................... | .................... | .................... | .................... | .................... | .................... |
| C | 5143 | hCoV-19/USA/MI-MDHHS-SC20448/2020\|EPI_ISL_452322\|2 | .................... | .................... | .................... | .................... | .................... | .................... |
| C | 5144 | hCoV-19/USA/MI-MDHHS-SC20449/2020\|EPI_ISL_452323\|2 | .................... | .................... | .................... | .................... | .................... | .................... |
| C | 5145 | hCoV-19/USA/MI-MDHHS-SC20450/2020\|EPI_ISL_452324\|2 | .................... | .................... | .................... | .................... | .................... | .................... |
| C | 5146 | hCoV-19/USA/MI-MDHHS-SC20451/2020\|EPI_ISL_452325\|2 | .................... | .................... | .................... | .................... | .................... | .................... |
| C | 5147 | hCoV-19/USA/MI-MDHHS-SC20452/2020\|EPI_ISL_452326\|2 | .................... | .................... | .................... | .................... | .................... | .................... |
| C | 5148 | hCoV-19/Beijing/DT-travelAT01/2020\|EPI_ISL_452327\| | .................... | .................... | .................... | .................... | .................... | .................... |
| C | 5149 | hCoV-19/Beijing/DT-travelPAK01/2020\|EPI_ISL_452328 | .................... | .................... | .................... | .................... | .................... | .................... |
| C | 5150 | hCoV-19/Beijing/DT-travelDE01/2020\|EPI_ISL_452329\| | .................... | .................... | .................... | .................... | .................... | .................... |
| C | 5151 | hCoV-19/Beijing/DT-travelFR01/2020\|EPI_ISL_452330\| | .................... | .................... | .................... | .................... | .................... | .................... |
| C | 5152 | hCoV-19/Beijing/DT-travelCA01/2020\|EPI_ISL_452331\| | .................... | .................... | .................... | .................... | .................... | .................... |
| C | 5153 | hCoV-19/Beijing/DT-travelUSA01/2020\|EPI_ISL_452332 | .................... | .................... | .................... | .................... | .................... | .................... |
| C | 5154 | hCoV-19/Beijing/DT-travelIRI01/2020\|EPI_ISL_452333 | .................... | .................... | .................... | .................... | .................... | .................... |
| C | 5155 | hCoV-19/Beijing/DT-travelIT05/2020\|EPI_ISL_452334\| | .................... | .................... | .................... | .................... | .................... | .................... |
| C | 5156 | hCoV-19/Beijing/DT-travelUK02/2020\|EPI_ISL_452335\| | .................... | .................... | .................... | .................... | .................... | .................... |
| C | 5157 | hCoV-19/Beijing/DT-travelUK04/2020\|EPI_ISL_452336\| | .................... | .................... | .................... | .................... | .................... | .................... |
| C | 5158 | hCoV-19/Beijing/DT-travelFR02/2020\|EPI_ISL_452337\| | .................... | .................... | .................... | .................... | .................... | .................... |
| C | 5159 | hCoV-19/Beijing/DT-travelUSA02/2020\|EPI_ISL_452338 | .................... | .................... | .................... | .................... | .................... | .................... |
| C | 5160 | hCoV-19/Beijing/DT-travelUSA04/2020\|EPI_ISL_452339 | .................... | .................... | .................... | .................... | .................... | .................... |
| C | 5161 | hCoV-19/Beijing/DT-travelUSA03/2020\|EPI_ISL_452340 | .................... | .................... | .................... | .................... | .................... | .................... |
| C | 5162 | hCoV-19/Beijing/DT-travelMX01/2020\|EPI_ISL_452341\| | .................... | .................... | .................... | .................... | .................... | .................... |
| C | 5163 | hCoV-19/Beijing/DT-travelNO01/2020\|EPI_ISL_452342\| | .................... | .................... | .................... | .................... | .................... | .................... |
| C | 5164 | hCoV-19/Beijing/DT-travelES01/2020\|EPI_ISL_452343\| | .................... | .................... | .................... | .................... | .................... | .................... |
| C | 5165 | hCoV-19/Beijing/DT-travelES02/2020\|EPI_ISL_452344\| | .................... | .................... | .................... | .................... | .................... | .................... |
| C | 5166 | hCoV-19/Beijing/DT-travelES03/2020\|EPI_ISL_452345\| | .................... | .................... | .................... | .................... | .................... | .................... |
| C | 5167 | hCoV-19/Beijing/DT-travelES04/2020\|EPI_ISL_452346\| | .................... | .................... | .................... | .................... | .................... | .................... |
| C | 5168 | hCoV-19/Beijing/DT-travelGR01/2020\|EPI_ISL_452347\| | .................... | .................... | .................... | .................... | .................... | .................... |
| C | 5169 | hCoV-19/Beijing/DT-travelHU02/2020\|EPI_ISL_452348\| | .................... | .................... | .................... | .................... | .................... | .................... |
| C | 5170 | hCoV-19/Beijing/DT-travelHU01/2020\|EPI_ISL_452349\| | .................... | .................... | .................... | .................... | .................... | .................... |
| C | 5171 | hCoV-19/Beijing/DT-travelHU03/2020\|EPI_ISL_452350\| | .................... | .................... | .................... | .................... | .................... | .................... |
| C | 5172 | hCoV-19/Beijing/DT-travelIT04/2020\|EPI_ISL_452351\| | .................... | .................... | .................... | .................... | .................... | .................... |
| C | 5173 | hCoV-19/Beijing/DT-travelIT01/2020\|EPI_ISL_452352\| | .................... | .................... | .................... | .................... | .................... | .................... |
| C | 5174 | hCoV-19/Beijing/DT-travelIT02/2020\|EPI_ISL_452353\| | .................... | .................... | .................... | .................... | .................... | .................... |
| C | 5175 | hCoV-19/Beijing/DT-travelIT03/2020\|EPI_ISL_452354\| | .................... | .................... | .................... | .................... | .................... | .................... |
| C | 5176 | hCoV-19/England/NORW-E94E3/2020\|EPI_ISL_453600\|202 | .................... | .................... | .................... | .................... | .................... | .................... |
| C | 5177 | hCoV-19/England/NORW-E9559/2020\|EPI_ISL_453601\|202 | .................... | .................... | .................... | .................... | .................... | .................... |
| C | 5178 | hCoV-19/England/NORW-E9595/2020\|EPI_ISL_453603\|202 | .................... | .................... | .................... | .................... | .................... | .................... |
| C | 5179 | hCoV-19/England/NORW-E95D1/2020\|EPI_ISL_453604\|202 | .................... | .................... | .................... | .................... | .................... | .................... |
| C | 5180 | hCoV-19/England/NORW-E95E0/2020\|EPI_ISL_453605\|202 | .................... | .................... | .................... | .................... | .................... | .................... |
| C | 5181 | hCoV-19/England/NORW-E95FF/2020\|EPI_ISL_453606\|202 | .................... | .................... | .................... | .................... | .................... | .................... |
| C | 5182 | hCoV-19/England/NORW-E9629/2020\|EPI_ISL_453607\|202 | .................... | .................... | .................... | .................... | .................... | .................... |
| C | 5183 | hCoV-19/England/NORW-E9638/2020\|EPI_ISL_453608\|202 | .................... | .................... | .................... | .................... | .................... | .................... |
| C | 5184 | hCoV-19/England/NORW-E9647/2020\|EPI_ISL_453609\|202 | .................... | .................... | .................... | .................... | .................... | .................... |
| C | 5185 | hCoV-19/England/NORW-E9674/2020\|EPI_ISL_453610\|202 | .................... | .................... | .................... | .................... | .................... | .................... |
| C | 5186 | hCoV-19/England/NORW-E96A1/2020\|EPI_ISL_453611\|202 | .................... | .................... | .................... | .................... | .................... | .................... |
| C | 5188 | hCoV-19/England/NORW-E96ED/2020\|EPI_ISL_453613\|202 | .................... | .................... | .................... | .................... | .................... | .................... |
| C | 5189 | hCoV-19/England/NOTT-11100B/2020\|EPI_ISL_453615\|20 | .................... | .................... | .................... | .................... | .................... | .................... |
| C | 5190 | hCoV-19/England/NOTT-11101A/2020\|EPI_ISL_453616\|20 | .................... | .................... | .................... | .................... | .................... | .................... |
| C | 5191 | hCoV-19/England/NOTT-111038/2020\|EPI_ISL_453618\|20 | .................... | .................... | .................... | .................... | .................... | .................... |
| C | 5192 | hCoV-19/England/NOTT-111047/2020\|EPI_ISL_453619\|20 | .................... | .................... | .................... | .................... | .................... | .................... |
| C | 5193 | hCoV-19/England/NOTT-111056/2020\|EPI_ISL_453620\|20 | .................... | .................... | .................... | .................... | .................... | .................... |
| C | 5194 | hCoV-19/England/NOTT-111065/2020\|EPI_ISL_453621\|20 | .................... | .................... | .................... | .................... | .................... | .................... |
| C | 5195 | hCoV-19/England/NOTT-111074/2020\|EPI_ISL_453622\|20 | .................... | .................... | .................... | .................... | .................... | .................... |
| C | 5196 | hCoV-19/England/NOTT-111083/2020\|EPI_ISL_453623\|20 | .................... | .................... | .................... | .................... | .................... | .................... |
| C | 5197 | hCoV-19/England/NOTT-111092/2020\|EPI_ISL_453624\|20 | .................... | .................... | .................... | .................... | .................... | .................... |
| C | 5198 | hCoV-19/England/NOTT-1110A1/2020\|EPI_ISL_453625\|20 | .................... | .................... | .................... | .................... | .................... | .................... |
| C | 5199 | hCoV-19/England/NOTT-1110B0/2020\|EPI_ISL_453626\|20 | .................... | .................... | .................... | .................... | .................... | .................... |
| C | 5200 | hCoV-19/England/NOTT-1110CF/2020\|EPI_ISL_453627\|20 | .................... | .................... | .................... | .................... | .................... | .................... |
| C | 5201 | hCoV-19/England/NOTT-1110DE/2020\|EPI_ISL_453628\|20 | .................... | .................... | .................... | .................... | .................... | .................... |
| C | 5202 | hCoV-19/England/NOTT-1110FC/2020\|EPI_ISL_453629\|20 | .................... | .................... | .................... | .................... | .................... | .................... |
| C | 5203 | hCoV-19/England/NOTT-111117/2020\|EPI_ISL_453631\|20 | .................... | .................... | .................... | .................... | .................... | .................... |
| C | 5204 | hCoV-19/England/NOTT-111126/2020\|EPI_ISL_453632\|20 | .................... | .................... | .................... | .................... | .................... | .................... |
| C | 5205 | hCoV-19/England/NOTT-111135/2020\|EPI_ISL_453633\|20 | .................... | .................... | .................... | .................... | .................... | .................... |
| C | 5206 | hCoV-19/England/NOTT-111144/2020\|EPI_ISL_453634\|20 | .................... | .................... | .................... | .................... | .................... | .................... |
| C | 5208 | hCoV-19/England/NOTT-11126F/2020\|EPI_ISL_453637\|20 | .................... | .................... | .................... | .................... | .................... | .................... |
| C | 5209 | hCoV-19/England/NOTT-11127E/2020\|EPI_ISL_453638\|20 | .................... | .................... | .................... | .................... | .................... | .................... |
| C | 5210 | hCoV-19/England/NOTT-11128D/2020\|EPI_ISL_453639\|20 | .................... | .................... | .................... | .................... | .................... | .................... |
| C | 5211 | hCoV-19/England/NOTT-11129C/2020\|EPI_ISL_453640\|20 | .................... | .................... | .................... | .................... | .................... | .................... |
| C | 5212 | hCoV-19/England/NOTT-1112AB/2020\|EPI_ISL_453641\|20 | .................... | .................... | .................... | .................... | .................... | .................... |
| C | 5213 | hCoV-19/England/NOTT-1112BA/2020\|EPI_ISL_453642\|20 | .................... | .................... | .................... | .................... | .................... | .................... |
| C | 5214 | hCoV-19/England/NOTT-1112E7/2020\|EPI_ISL_453645\|20 | .................... | .................... | .................... | .................... | .................... | .................... |
| C | 5215 | hCoV-19/England/NOTT-1112F6/2020\|EPI_ISL_453646\|20 | .................... | .................... | .................... | .................... | .................... | .................... |
| C | 5216 | hCoV-19/England/NOTT-111302/2020\|EPI_ISL_453647\|20 | .................... | .................... | .................... | .................... | .................... | .................... |
| C | 5217 | hCoV-19/England/NOTT-11133F/2020\|EPI_ISL_453648\|20 | .................... | .................... | .................... | .................... | .................... | .................... |
| C | 5218 | hCoV-19/England/NOTT-11134E/2020\|EPI_ISL_453649\|20 | .................... | .................... | .................... | .................... | .................... | .................... |
| C | 5219 | hCoV-19/England/NOTT-11135D/2020\|EPI_ISL_453650\|20 | .................... | .................... | .................... | .................... | .................... | .................... |
| C | 5220 | hCoV-19/England/NOTT-11136C/2020\|EPI_ISL_453651\|20 | .................... | .................... | .................... | .................... | .................... | .................... |

| | | | | | | | | |
|---|---|---|---|---|---|---|---|---|
| C | 5221 | hCoV-19/England/NOTT-11137B/2020\|EPI_ISL_453652\|20 | .................... | .................... | .................... | .................... | .................... | .................... |
| C | 5222 | hCoV-19/England/NOTT-1113A8/2020\|EPI_ISL_453655\|20 | .................... | .................... | .................... | .................... | .................... | .................... |
| C | 5223 | hCoV-19/England/NOTT-1113B7/2020\|EPI_ISL_453656\|20 | .................... | .................... | .................... | .................... | .................... | .................... |
| C | 5224 | hCoV-19/England/NOTT-1113C6/2020\|EPI_ISL_453657\|20 | .................... | .................... | .................... | .................... | .................... | .................... |
| C | 5225 | hCoV-19/England/NOTT-1113D5/2020\|EPI_ISL_453658\|20 | .................... | .................... | .................... | .................... | .................... | .................... |
| C | 5226 | hCoV-19/England/NOTT-1113E4/2020\|EPI_ISL_453659\|20 | .................... | .................... | .................... | .................... | .................... | .................... |
| C | 5227 | hCoV-19/England/NOTT-11141E/2020\|EPI_ISL_453662\|20 | .................... | .................... | .................... | .................... | .................... | .................... |
| C | 5228 | hCoV-19/England/NOTT-11142D/2020\|EPI_ISL_453663\|20 | .................... | .................... | .................... | .................... | .................... | .................... |
| C | 5229 | hCoV-19/England/PORT-2CF302/2020\|EPI_ISL_453670\|20 | .................... | .................... | .................... | .................... | .................... | .................... |
| C | 5230 | hCoV-19/England/PORT-2CF3E4/2020\|EPI_ISL_453676\|20 | .................... | .................... | .................... | .................... | .................... | .................... |
| C | 5231 | hCoV-19/England/PORT-2CF3F3/2020\|EPI_ISL_453677\|20 | .................... | .................... | .................... | .................... | .................... | .................... |
| C | 5232 | hCoV-19/England/PORT-2CF42D/2020\|EPI_ISL_453679\|20 | .................... | .................... | .................... | .................... | .................... | .................... |
| C | 5233 | hCoV-19/England/PORT-2CF496/2020\|EPI_ISL_453684\|20 | .................... | .................... | .................... | .................... | .................... | .................... |
| C | 5234 | hCoV-19/England/PORT-2CF4A5/2020\|EPI_ISL_453685\|20 | .................... | .................... | .................... | .................... | .................... | .................... |
| C | 5235 | hCoV-19/Wuhan/HB-WH2-163/2020\|EPI_ISL_454930\|2020- | .................... | .................... | .................... | .................... | .................... | .................... |
| C | 5236 | hCoV-19/Wuhan/HB-WH2-164/2020\|EPI_ISL_454931\|2020- | .................... | .................... | .................... | .................... | .................... | .................... |
| C | 5237 | hCoV-19/Wuhan/HB-WH2-166/2020\|EPI_ISL_454932\|2020- | .................... | .................... | .................... | .................... | .................... | .................... |
| C | 5238 | hCoV-19/Wuhan/HB-WH2-167/2020\|EPI_ISL_454933\|2020- | .................... | .................... | .................... | .................... | .................... | .................... |
| C | 5239 | hCoV-19/Wuhan/HB-WH2-168/2020\|EPI_ISL_454934\|2020- | .................... | .................... | .................... | .................... | .................... | .................... |
| C | 5240 | hCoV-19/Wuhan/HB-WH2-169/2020\|EPI_ISL_454935\|2020- | .................... | .................... | .................... | .................... | .................... | .................... |
| C | 5241 | hCoV-19/Wuhan/HB-WH3-170/2020\|EPI_ISL_454936\|2020- | .................... | .................... | .................... | .................... | .................... | .................... |
| C | 5242 | hCoV-19/Wuhan/HB-WH3-171/2020\|EPI_ISL_454937\|2020- | .................... | .................... | .................... | .................... | .................... | .................... |
| C | 5243 | hCoV-19/Wuhan/HB-WH3-172/2020\|EPI_ISL_454938\|2020- | .................... | .................... | .................... | .................... | .................... | .................... |
| C | 5244 | hCoV-19/Wuhan/HB-WH3-173/2020\|EPI_ISL_454939\|2020- | .................... | .................... | .................... | .................... | .................... | .................... |
| C | 5245 | hCoV-19/Wuhan/HB-WH3-176/2020\|EPI_ISL_454940\|2020- | .................... | .................... | .................... | .................... | .................... | .................... |
| C | 5246 | hCoV-19/Wuhan/HB-WH3-179/2020\|EPI_ISL_454941\|2020- | .................... | .................... | .................... | .................... | .................... | .................... |
| C | 5247 | hCoV-19/Wuhan/HB-WH3-181/2020\|EPI_ISL_454942\|2020- | .................... | .................... | .................... | .................... | .................... | .................... |
| C | 5248 | hCoV-19/Wuhan/HB-WH3-182/2020\|EPI_ISL_454943\|2020- | .................... | .................... | .................... | .................... | .................... | .................... |
| C | 5249 | hCoV-19/Wuhan/HB-WH3-183/2020\|EPI_ISL_454944\|2020- | .................... | .................... | .................... | .................... | .................... | .................... |
| C | 5250 | hCoV-19/Wuhan/HB-WH3-189/2020\|EPI_ISL_454946\|2020- | .................... | .................... | .................... | .................... | .................... | .................... |
| C | 5251 | hCoV-19/Wuhan/HB-WH3-191/2020\|EPI_ISL_454947\|2020- | .................... | .................... | .................... | .................... | .................... | .................... |
| C | 5252 | hCoV-19/Wuhan/HB-WH4-194/2020\|EPI_ISL_454948\|2020- | .................... | .................... | .................... | .................... | .................... | .................... |
| C | 5253 | hCoV-19/Wuhan/HB-WH4-195/2020\|EPI_ISL_454949\|2020- | .................... | .................... | .................... | .................... | .................... | .................... |
| C | 5254 | hCoV-19/Wuhan/HB-WH4-196/2020\|EPI_ISL_454950\|2020- | .................... | .................... | .................... | .................... | .................... | .................... |
| C | 5255 | hCoV-19/Wuhan/HB-WH4-197/2020\|EPI_ISL_454951\|2020- | .................... | .................... | .................... | .................... | .................... | .................... |
| C | 5256 | hCoV-19/Wuhan/HB-WH4-200/2020\|EPI_ISL_454952\|2020- | .................... | .................... | .................... | .................... | .................... | .................... |
| C | 5257 | hCoV-19/Wuhan/HB-WH4-201/2020\|EPI_ISL_454953\|2020- | .................... | .................... | .................... | .................... | .................... | .................... |
| C | 5258 | hCoV-19/Wuhan/HB-WH4-204/2020\|EPI_ISL_454954\|2020- | .................... | .................... | .................... | .................... | .................... | .................... |
| C | 5259 | hCoV-19/Wuhan/HB-WH4-205/2020\|EPI_ISL_454955\|2020- | .................... | .................... | .................... | .................... | .................... | .................... |
| C | 5260 | hCoV-19/Wuhan/HB-WH4-206/2020\|EPI_ISL_454956\|2020- | .................... | .................... | .................... | .................... | .................... | .................... |
| C | 5261 | hCoV-19/Wuhan/HB-WH4-208/2020\|EPI_ISL_454957\|2020- | .................... | .................... | .................... | .................... | .................... | .................... |
| C | 5262 | hCoV-19/Wuhan/HB-WH4-209/2020\|EPI_ISL_454958\|2020- | .................... | .................... | .................... | .................... | .................... | .................... |
| C | 5263 | hCoV-19/Wuhan/HB-WH4-211/2020\|EPI_ISL_454960\|2020- | .................... | .................... | .................... | .................... | .................... | .................... |
| C | 5264 | hCoV-19/Wuhan/HB-WH4-212/2020\|EPI_ISL_454961\|2020- | .................... | .................... | .................... | .................... | .................... | .................... |
| C | 5265 | hCoV-19/Wuhan/HB-WH4-213/2020\|EPI_ISL_454962\|2020- | .................... | .................... | .................... | .................... | .................... | .................... |
| C | 5266 | hCoV-19/Wuhan/HB-WH4-214/2020\|EPI_ISL_454963\|2020- | .................... | .................... | .................... | .................... | .................... | .................... |
| C | 5267 | hCoV-19/Wuhan/HB-WH5-220/2020\|EPI_ISL_454965\|2020- | .................... | .................... | .................... | .................... | .................... | .................... |
| C | 5268 | hCoV-19/Wuhan/HB-WH5-222/2020\|EPI_ISL_454967\|2020- | .................... | .................... | .................... | .................... | .................... | .................... |
| C | 5269 | hCoV-19/Wuhan/HB-WH5-223/2020\|EPI_ISL_454968\|2020- | .................... | .................... | .................... | .................... | .................... | .................... |
| C | 5270 | hCoV-19/Wuhan/HB-WH5-224/2020\|EPI_ISL_454969\|2020- | .................... | .................... | .................... | .................... | .................... | .................... |
| C | 5271 | hCoV-19/Wuhan/HB-WH5-226/2020\|EPI_ISL_454971\|2020- | .................... | .................... | .................... | .................... | .................... | .................... |
| C | 5272 | hCoV-19/Wuhan/HB-WH5-227/2020\|EPI_ISL_454972\|2020- | .................... | .................... | .................... | .................... | .................... | .................... |
| C | 5273 | hCoV-19/Wuhan/HB-WH5-229/2020\|EPI_ISL_454973\|2020- | .................... | .................... | .................... | .................... | .................... | .................... |
| C | 5274 | hCoV-19/Wuhan/HB-WH5-231/2020\|EPI_ISL_454974\|2020- | .................... | .................... | .................... | .................... | .................... | .................... |
| C | 5275 | hCoV-19/Wuhan/HB-WH5-232/2020\|EPI_ISL_454975\|2020- | .................... | .................... | .................... | .................... | .................... | .................... |
| C | 5276 | hCoV-19/Wuhan/HB-WH5-233/2020\|EPI_ISL_454976\|2020- | .................... | .................... | .................... | .................... | .................... | .................... |
| C | 5277 | hCoV-19/Wuhan/HB-WH5-236/2020\|EPI_ISL_454977\|2020- | .................... | .................... | .................... | .................... | .................... | .................... |
| C | 5278 | hCoV-19/Wuhan/HB-WH5-237/2020\|EPI_ISL_454978\|2020- | .................... | .................... | .................... | .................... | .................... | .................... |
| C | 5279 | hCoV-19/Wuhan/HB-WH5-238/2020\|EPI_ISL_454979\|2020- | .................... | .................... | .................... | .................... | .................... | .................... |
| C | 5280 | hCoV-19/Wuhan/HB-WH5-239/2020\|EPI_ISL_454980\|2020- | .................... | .................... | .................... | .................... | .................... | .................... |
| C | 5281 | hCoV-19/Wuhan/HB-WH5-240/2020\|EPI_ISL_454981\|2020- | .................... | .................... | .................... | .................... | .................... | .................... |
| C | 5282 | hCoV-19/Wuhan/HB-WH6-242/2020\|EPI_ISL_454982\|2020- | .................... | .................... | .................... | .................... | .................... | .................... |
| C | 5283 | hCoV-19/Wuhan/HB-WH6-246/2020\|EPI_ISL_454983\|2020- | .................... | .................... | .................... | .................... | .................... | .................... |
| C | 5284 | hCoV-19/Wuhan/HB-WH6-247/2020\|EPI_ISL_454984\|2020- | .................... | .................... | .................... | .................... | .................... | .................... |
| C | 5285 | hCoV-19/Wuhan/HB-WHCM-102/2020\|EPI_ISL_454988\|2020 | .................... | .................... | .................... | .................... | .................... | .................... |
| C | 5286 | hCoV-19/Wuhan/HB-WHCM-104/2020\|EPI_ISL_454989\|2020 | .................... | .................... | .................... | .................... | .................... | .................... |
| C | 5287 | hCoV-19/Wuhan/HB-WHCM-107/2020\|EPI_ISL_454990\|2020 | .................... | .................... | .................... | .................... | .................... | .................... |
| C | 5288 | hCoV-19/Wuhan/HB-WHCM-108/2020\|EPI_ISL_454991\|2020 | .................... | .................... | .................... | .................... | .................... | .................... |
| C | 5289 | hCoV-19/Wuhan/HB-WHCM-109/2020\|EPI_ISL_454992\|2020 | .................... | .................... | .................... | .................... | .................... | .................... |
| C | 5290 | hCoV-19/Wuhan/HB-WHCM-113/2020\|EPI_ISL_454993\|2020 | .................... | .................... | .................... | .................... | .................... | .................... |
| C | 5291 | hCoV-19/Wuhan/HB-WHCM-115/2020\|EPI_ISL_454995\|2020 | .................... | .................... | .................... | .................... | .................... | .................... |
| C | 5292 | hCoV-19/Wuhan/HB-WHCM-116/2020\|EPI_ISL_454996\|2020 | .................... | .................... | .................... | .................... | .................... | .................... |
| C | 5293 | hCoV-19/Wuhan/HB-WHCM-117/2020\|EPI_ISL_454997\|2020 | .................... | .................... | .................... | .................... | .................... | .................... |
| C | 5294 | hCoV-19/England/EXET-135A5E/2020\|EPI_ISL_457129\|20 | .................... | .................... | .................... | .................... | .................... | .................... |
| C | 5295 | hCoV-19/England/EXET-135AA9/2020\|EPI_ISL_457132\|20 | .................... | .................... | .................... | .................... | .................... | .................... |
| C | 5296 | hCoV-19/England/EXET-135AE5/2020\|EPI_ISL_457133\|20 | .................... | .................... | .................... | .................... | .................... | .................... |
| C | 5297 | hCoV-19/England/EXET-135B00/2020\|EPI_ISL_457134\|20 | .................... | .................... | .................... | .................... | .................... | .................... |
| C | 5298 | hCoV-19/England/EXET-135B2E/2020\|EPI_ISL_457135\|20 | .................... | .................... | .................... | .................... | .................... | .................... |

| | | | | | | | |
|---|---|---|---|---|---|---|---|
| C | 5299 hCoV-19/England/EXET-135B88/2020\|EPI_ISL_457139\|20 | ···················· | ···················· | ····················· | ···················· | ···················· | ···················· |
| C | 5300 hCoV-19/England/EXET-135B97/2020\|EPI_ISL_457140\|20 | ···················· | ···················· | ····················· | ···················· | ···················· | ···················· |
| C | 5301 hCoV-19/England/EXET-135BA6/2020\|EPI_ISL_457141\|20 | ···················· | ···················· | ····················· | ···················· | ···················· | ···················· |
| C | 5302 hCoV-19/England/EXET-135BB5/2020\|EPI_ISL_457142\|20 | ···················· | ···················· | ····················· | ···················· | ···················· | ···················· |
| C | 5303 hCoV-19/England/EXET-135BC4/2020\|EPI_ISL_457143\|20 | ···················· | ···················· | ····················· | ···················· | ···················· | ···················· |
| C | 5304 hCoV-19/England/EXET-135BD3/2020\|EPI_ISL_457144\|20 | ···················· | ···················· | ····················· | ···················· | ···················· | ···················· |
| C | 5305 hCoV-19/England/EXET-135BE2/2020\|EPI_ISL_457145\|20 | ···················· | ···················· | ····················· | ···················· | ···················· | ···················· |
| C | 5306 hCoV-19/England/EXET-135C0D/2020\|EPI_ISL_457146\|20 | ···················· | ···················· | ····················· | ···················· | ···················· | ···················· |
| C | 5307 hCoV-19/England/EXET-135C1C/2020\|EPI_ISL_457147\|20 | ···················· | ···················· | ····················· | ···················· | ···················· | ···················· |
| C | 5308 hCoV-19/England/EXET-135C2B/2020\|EPI_ISL_457148\|20 | ···················· | ···················· | ····················· | ···················· | ···················· | ···················· |
| C | 5309 hCoV-19/England/EXET-135C3A/2020\|EPI_ISL_457149\|20 | ···················· | ···················· | ····················· | ···················· | ···················· | ···················· |
| C | 5310 hCoV-19/England/EXET-135C76/2020\|EPI_ISL_457150\|20 | ···················· | ···················· | ····················· | ···················· | ···················· | ···················· |
| C | 5311 hCoV-19/England/EXET-135C85/2020\|EPI_ISL_457151\|20 | ···················· | ···················· | ····················· | ···················· | ···················· | ···················· |
| C | 5312 hCoV-19/England/EXET-135C94/2020\|EPI_ISL_457152\|20 | ···················· | ···················· | ····················· | ···················· | ···················· | ···················· |
| C | 5313 hCoV-19/England/EXET-135CA3/2020\|EPI_ISL_457153\|20 | ···················· | ···················· | ····················· | ···················· | ···················· | ···················· |
| C | 5314 hCoV-19/England/EXET-135CC1/2020\|EPI_ISL_457154\|20 | ···················· | ···················· | ····················· | ···················· | ···················· | ···················· |
| C | 5315 hCoV-19/England/EXET-135CD0/2020\|EPI_ISL_457155\|20 | ···················· | ···················· | ····················· | ···················· | ···················· | ···················· |
| C | 5316 hCoV-19/England/EXET-135CEF/2020\|EPI_ISL_457156\|20 | ···················· | ···················· | ····················· | ···················· | ···················· | ···················· |
| C | 5317 hCoV-19/England/EXET-135D0A/2020\|EPI_ISL_457157\|20 | ···················· | ···················· | ····················· | ···················· | ···················· | ···················· |
| C | 5318 hCoV-19/England/EXET-135D28/2020\|EPI_ISL_457159\|20 | ···················· | ···················· | ····················· | ···················· | ···················· | ···················· |
| C | 5319 hCoV-19/England/EXET-135D37/2020\|EPI_ISL_457160\|20 | ···················· | ···················· | ····················· | ···················· | ···················· | ···················· |
| C | 5320 hCoV-19/England/EXET-135D46/2020\|EPI_ISL_457161\|20 | ···················· | ···················· | ····················· | ···················· | ···················· | ···················· |
| C | 5321 hCoV-19/England/EXET-135D55/2020\|EPI_ISL_457162\|20 | ···················· | ···················· | ····················· | ···················· | ···················· | ···················· |
| C | 5322 hCoV-19/England/EXET-135D64/2020\|EPI_ISL_457163\|20 | ···················· | ···················· | ····················· | ···················· | ···················· | ···················· |
| C | 5323 hCoV-19/England/EXET-135D91/2020\|EPI_ISL_457164\|20 | ···················· | ···················· | ····················· | ···················· | ···················· | ···················· |
| C | 5324 hCoV-19/England/EXET-135DA0/2020\|EPI_ISL_457165\|20 | ···················· | ···················· | ····················· | ···················· | ···················· | ···················· |
| C | 5325 hCoV-19/England/EXET-135DBF/2020\|EPI_ISL_457166\|20 | ···················· | ···················· | ····················· | ···················· | ···················· | ···················· |
| C | 5326 hCoV-19/England/EXET-135DCE/2020\|EPI_ISL_457167\|20 | ···················· | ···················· | ····················· | ···················· | ···················· | ···················· |
| C | 5327 hCoV-19/England/EXET-135DDD/2020\|EPI_ISL_457168\|20 | ···················· | ···················· | ····················· | ···················· | ···················· | ···················· |
| C | 5328 hCoV-19/England/EXET-135DEC/2020\|EPI_ISL_457169\|20 | ···················· | ···················· | ····················· | ···················· | ···················· | ···················· |
| C | 5329 hCoV-19/England/BRIS-12A737/2020\|EPI_ISL_458465\|20 | ···················· | ···················· | ····················· | ···················· | ···················· | ···················· |
| C | 5330 hCoV-19/England/BRIS-12996F/2020\|EPI_ISL_458466\|20 | ···················· | ···················· | ····················· | ···················· | ···················· | ···················· |
| C | 5331 hCoV-19/England/BRIS-129ECA/2020\|EPI_ISL_458468\|20 | ···················· | ···················· | ····················· | ···················· | ···················· | ···················· |
| C | 5332 hCoV-19/England/BRIS-12A685/2020\|EPI_ISL_458469\|20 | ···················· | ···················· | ····················· | ···················· | ···················· | ···················· |
| C | 5333 hCoV-19/England/BRIS-12AEE7/2020\|EPI_ISL_458470\|20 | ···················· | ··············Y····· | ····················· | ···················· | ···················· | ···················· |
| C | 5334 hCoV-19/England/BRIS-12AA10/2020\|EPI_ISL_458471\|20 | ···················· | ···················· | ····················· | ···················· | ···················· | ···················· |
| C | 5335 hCoV-19/England/BRIS-12A4E5/2020\|EPI_ISL_458472\|20 | ···················· | ···················· | ····················· | ···················· | ···················· | ···················· |
| C | 5336 hCoV-19/England/BRIS-12A834/2020\|EPI_ISL_458474\|20 | ···················· | ···················· | ····················· | ···················· | ···················· | ···················· |
| C | 5337 hCoV-19/England/BRIS-12A7EC/2020\|EPI_ISL_458476\|20 | ···················· | ···················· | ····················· | ···················· | ···················· | ···················· |
| C | 5338 hCoV-19/England/BRIS-12AE7E/2020\|EPI_ISL_458479\|20 | ···················· | ···················· | ····················· | ···················· | ···················· | ···················· |
| C | 5339 hCoV-19/England/BRIS-12A096/2020\|EPI_ISL_458480\|20 | ···················· | ···················· | ····················· | ···················· | ···················· | ···················· |
| C | 5340 hCoV-19/England/BRIS-12A791/2020\|EPI_ISL_458483\|20 | ···················· | ···················· | ····················· | ···················· | ···················· | ···················· |
| C | 5341 hCoV-19/England/BRIS-12A95F/2020\|EPI_ISL_458484\|20 | ···················· | ···················· | ····················· | ···················· | ···················· | ···················· |
| C | 5342 hCoV-19/England/BRIS-12A6FE/2020\|EPI_ISL_458485\|20 | ···················· | ···················· | ····················· | ···················· | ···················· | ···················· |
| C | 5343 hCoV-19/England/BRIS-12AF4E/2020\|EPI_ISL_458486\|20 | ···················· | ···················· | ····················· | ···················· | ···················· | ···················· |
| C | 5344 hCoV-19/England/BRIS-12A70A/2020\|EPI_ISL_458489\|20 | ···················· | ···················· | ····················· | ···················· | ···················· | ···················· |
| C | 5345 hCoV-19/England/BRIS-12AA2F/2020\|EPI_ISL_458490\|20 | ···················· | ···················· | ····················· | ···················· | ···················· | ···················· |
| C | 5346 hCoV-19/England/BRIS-12A53D/2020\|EPI_ISL_458492\|20 | ···················· | ···················· | ····················· | ···················· | ···················· | ···················· |
| C | 5347 hCoV-19/England/BRIS-12A49A/2020\|EPI_ISL_458495\|20 | ···················· | ···················· | ····················· | ···················· | ···················· | ···················· |
| C | 5348 hCoV-19/England/BRIS-12AD35/2020\|EPI_ISL_458496\|20 | ···················· | ···················· | ····················· | ···················· | ···················· | ···················· |
| C | 5349 hCoV-19/England/BRIS-12A807/2020\|EPI_ISL_458497\|20 | ···················· | ···················· | ····················· | ···················· | ···················· | ···················· |
| C | 5350 hCoV-19/Netherlands/NA_607/2020\|EPI_ISL_460770\|202 | ···················· | ···················· | ····················· | ···················· | ···················· | ···················· |
| C | 5351 hCoV-19/Netherlands/NA_609/2020\|EPI_ISL_460772\|202 | ···················· | ···················· | ····················· | ···················· | ···················· | ···················· |
| C | 5352 hCoV-19/Netherlands/NA_610/2020\|EPI_ISL_460773\|202 | ···················· | ···················· | ····················· | ···················· | ···················· | ···················· |
| C | 5353 hCoV-19/Netherlands/NA_611/2020\|EPI_ISL_460774\|202 | ···················· | ···················· | ····················· | ···················· | ···················· | ···················· |
| C | 5354 hCoV-19/Netherlands/NA_612/2020\|EPI_ISL_460775\|202 | ···················· | ···················· | ····················· | ···················· | ···················· | ···················· |
| C | 5355 hCoV-19/Netherlands/NA_613/2020\|EPI_ISL_460776\|202 | ···················· | ···················· | ····················· | ···················· | ···················· | ···················· |
| C | 5356 hCoV-19/Netherlands/NA_614/2020\|EPI_ISL_460777\|202 | ···················· | ···················· | ····················· | ···················· | ···················· | ···················· |
| C | 5357 hCoV-19/Netherlands/NA_617/2020\|EPI_ISL_460780\|202 | ···················· | ···················· | ····················· | ···················· | ···················· | ···················· |
| C | 5358 hCoV-19/Netherlands/NA_618/2020\|EPI_ISL_460781\|202 | ···················· | ···················· | ····················· | ···················· | ···················· | ···················· |
| C | 5359 hCoV-19/Netherlands/NA_619/2020\|EPI_ISL_460782\|202 | ···················· | ···················· | ····················· | ···················· | ···················· | ···················· |
| C | 5360 hCoV-19/Netherlands/NA_622/2020\|EPI_ISL_460785\|202 | ···················· | ···········A········ | ····················· | ···················· | ···················· | ···················· |
| C | 5361 hCoV-19/Netherlands/NA_623/2020\|EPI_ISL_460786\|202 | ···················· | ···················· | ····················· | ···················· | ···················· | ···················· |
| C | 5362 hCoV-19/Netherlands/NA_624/2020\|EPI_ISL_460787\|202 | ···················· | ···················· | ····················· | ···················· | ···················· | ···················· |
| C | 5363 hCoV-19/Netherlands/NA_625/2020\|EPI_ISL_460788\|202 | ···················· | ···················· | ····················· | ···················· | ···················· | ···················· |
| C | 5364 hCoV-19/Netherlands/NoordBrabant_108/2020\|EPI_ISL_ | ···················· | ···················· | ····················· | ···················· | ···················· | ···················· |
| C | 5365 hCoV-19/Netherlands/NoordBrabant_109/2020\|EPI_ISL_ | ···················· | ···················· | ····················· | ···················· | ···················· | ···················· |
| C | 5366 hCoV-19/Netherlands/NoordHolland_10/2020\|EPI_ISL_4 | ···················· | ···················· | ····················· | ···················· | ···················· | ···················· |
| C | 5367 hCoV-19/Netherlands/NoordHolland_11/2020\|EPI_ISL_4 | ···················· | ···················· | ····················· | ···················· | ···················· | ···················· |
| C | 5368 hCoV-19/Netherlands/NoordHolland_12/2020\|EPI_ISL_4 | ···················· | ···················· | ····················· | ···················· | ···················· | ···················· |
| C | 5369 hCoV-19/Netherlands/NoordHolland_13/2020\|EPI_ISL_4 | ···················· | ···················· | ····················· | ···················· | ···················· | ···················· |
| C | 5370 hCoV-19/Netherlands/NoordHolland_6/2020\|EPI_ISL_46 | ···················· | ···················· | ····················· | ···················· | ···················· | ···················· |
| C | 5371 hCoV-19/Netherlands/NoordHolland_8/2020\|EPI_ISL_46 | ···················· | ···················· | ····················· | ···················· | ···················· | ···················· |
| C | 5372 hCoV-19/Netherlands/NoordHolland_9/2020\|EPI_ISL_46 | ···················· | ···················· | ····················· | ···················· | ···················· | ···················· |
| C | 5373 hCoV-19/Netherlands/Overijssel_10/2020\|EPI_ISL_460 | ···················· | ···················· | ····················· | ···················· | ···················· | ···················· |
| C | 5374 hCoV-19/Spain/AS-MG-100-20/2020\|EPI_ISL_463000\|202 | ···················· | ···················· | ····················· | ···················· | ···················· | ···················· |
| C | 5375 hCoV-19/Tunisia/COV0010-12/2020\|EPI_ISL_463001\|202 | ···················· | ···················· | ····················· | ···················· | ···················· | ···················· |
| C | 5376 hCoV-19/Tunisia/COV0880/2020\|EPI_ISL_463002\|2020-0 | ···················· | ···················· | ····················· | ···················· | ···················· | ···················· |

C 5377 hCoV-19/Colombia/CAL-GUR-0072/2020|EPI_ISL_447734| .................... .................... .................... .................... .................... ....................
C 5378 hCoV-19/Colombia/VAC-GUR-0084/2020|EPI_ISL_447735| .................... .................... .................... .................... .................... ....................
C 5379 hCoV-19/Colombia/VAC-GUR-0094/2020|EPI_ISL_447736| .................... .................... .................... .................... .................... ....................
C 5380 hCoV-19/Colombia/BOY-GUR-0206/2020|EPI_ISL_447738| .................... .................... .................... .................... .................... ....................
C 5381 hCoV-19/Colombia/BOY-GUR-0207/2020|EPI_ISL_447739| .................... .................... .................... .................... .................... ....................
C 5382 hCoV-19/Colombia/BOY-GUR-0209/2020|EPI_ISL_447740| .................... .................... .................... .................... .................... ....................
C 5383 hCoV-19/Colombia/BOY-GUR-0210/2020|EPI_ISL_447741| .................... .................... .................... .................... .................... ....................
C 5384 hCoV-19/Colombia/BOY-GUR-0212/2020|EPI_ISL_447742| .................... .................... .................... .................... .................... ....................
C 5385 hCoV-19/Colombia/BOY-GUR-0215/2020|EPI_ISL_447743| .................... .................... .................... .................... .................... ....................
C 5386 hCoV-19/Colombia/CUN-GUR-0454/2020|EPI_ISL_447744| .................... .................... .................... .................... .................... ....................
C 5387 hCoV-19/Colombia/CUN-GUR-0458/2020|EPI_ISL_447745| .................... .................... .................... .................... .................... ....................
C 5388 hCoV-19/Colombia/CUN-GUR-0459/2020|EPI_ISL_447746| .................... .................... .................... .................... .................... ....................
C 5389 hCoV-19/Colombia/BOY-GUR-0574/2020|EPI_ISL_447747| .................... .................... .................... .................... .................... ....................
C 5390 hCoV-19/Colombia/NAR-GUR-0609/2020|EPI_ISL_447748| .................... .................... .................... .................... .................... ....................
C 5391 hCoV-19/Colombia/CHO-GUR-0787/2020|EPI_ISL_447750| .................... .................... .................... .................... .................... ....................
C 5392 hCoV-19/Colombia/CUN-GUR-1215/2020|EPI_ISL_447754| .................... .................... .................... .................... .................... ....................
C 5393 hCoV-19/Colombia/VAC-GUV-92034/2020|EPI_ISL_447755 .................... .................... .................... .................... .................... ....................
C 5394 hCoV-19/Colombia/VAC-GUV-92042/2020|EPI_ISL_447756 .................... .................... .................... .................... .................... ....................
C 5395 hCoV-19/Colombia/TOL-GUV-92052/2020|EPI_ISL_447757 .................... .................... .................... .................... .................... ....................
C 5396 hCoV-19/Colombia/HUI-GUV-92061/2020|EPI_ISL_447759 .................... .................... .................... .................... .................... ....................
C 5397 hCoV-19/Colombia/RIS-GUV-92087/2020|EPI_ISL_447760 .................... .................... .................... .................... .................... ....................
C 5398 hCoV-19/Colombia/ANT-GUV-92146/2020|EPI_ISL_447761 .................... .................... .................... .................... .................... ....................
C 5399 hCoV-19/Colombia/ANT-GUV-92155/2020|EPI_ISL_447762 .................... .................... .................... .................... .................... ....................
C 5400 hCoV-19/Colombia/ANT-GVI-92161/2020|EPI_ISL_447763 .................... .................... .................... .................... .................... ....................
C 5401 hCoV-19/Colombia/ANT-GUV-92218/2020|EPI_ISL_447765 .................... .................... .................... .................... .................... ....................
C 5402 hCoV-19/Colombia/ANT-GUV-92250/2020|EPI_ISL_447766 .................... .................... .................... .................... .................... ....................
C 5403 hCoV-19/Colombia/ANT-GUV-92252/2020|EPI_ISL_447767 .................... .................... .................... .................... .................... ....................
C 5404 hCoV-19/Colombia/ANT-GUV-92256/2020|EPI_ISL_447768 .................... .................... .................... .................... .................... ....................
C 5406 hCoV-19/Colombia/VAC-GVI-93352/2020|EPI_ISL_447771 .................... .................... .................... .................... .................... ....................
C 5407 hCoV-19/Colombia/VAC-GVI-93366/2020|EPI_ISL_447774 .................... .................... .................... .................... .................... ....................
C 5408 hCoV-19/Colombia/VAC-GVI-93381/2020|EPI_ISL_447775 .................... .................... .................... .................... .................... ....................
C 5409 hCoV-19/Colombia/MAG-GVI-93450/2020|EPI_ISL_447777 .................... .................... .................... .................... .................... ....................
C 5410 hCoV-19/Colombia/MAG-GVI-93457/2020|EPI_ISL_447778 .................... .................... .................... .................... .................... ....................
C 5411 hCoV-19/Colombia/VAC-GVI-93467/2020|EPI_ISL_447779 .................... .................... .................... .................... .................... ....................
C 5412 hCoV-19/Colombia/VAC-GVI-93470/2020|EPI_ISL_447780 .................... .................... .................... .................... .................... ....................
C 5413 hCoV-19/Colombia/VAC-GVI-93473/2020|EPI_ISL_447781 .................... .................... .................... .................... .................... ....................
C 5414 hCoV-19/Colombia/VAC-GVI-93511/2020|EPI_ISL_447785 .................... .................... .................... .................... .................... ....................
C 5415 hCoV-19/Colombia/VAC-GVI-93534/2020|EPI_ISL_447786 .................... .................... .................... .................... .................... ....................
C 5416 hCoV-19/Colombia/VAC-GVI-93537/2020|EPI_ISL_447789 .................... .................... .................... .................... .................... ....................
C 5417 hCoV-19/Colombia/VAC-GVI-93538/2020|EPI_ISL_447790 .................... .................... .................... .................... .................... ....................
C 5418 hCoV-19/Colombia/VAC-GVI-97161/2020|EPI_ISL_447791 .................... .................... .................... .................... .................... ....................
C 5419 hCoV-19/Colombia/VAC-GVI-97181/2020|EPI_ISL_447792 .................... .................... .................... .................... .................... ....................
C 5420 hCoV-19/Colombia/VAC-GVI-97203/2020|EPI_ISL_447793 .................... .................... .................... .................... .................... ....................
C 5421 hCoV-19/Colombia/VAC-GVI-97210/2020|EPI_ISL_447794 .................... .................... .................... .................... .................... ....................
C 5422 hCoV-19/Colombia/VAC-GVI-97212/2020|EPI_ISL_447795 .................... .................... .................... .................... .................... ....................
C 5423 hCoV-19/Colombia/VAC-GVI-97321/2020|EPI_ISL_447796 .................... .................... .................... .................... .................... ....................
C 5424 hCoV-19/Colombia/BOY-GVI-97349/2020|EPI_ISL_447797 .................... .................... .................... .................... .................... ....................
C 5425 hCoV-19/Colombia/NAR-GVI-97417/2020|EPI_ISL_447798 .................... .................... .................... .................... .................... ....................
C 5426 hCoV-19/USA/FL-UF9/2020|EPI_ISL_447905|2020-04-29 .................... .................... .................... .................... .................... ....................
C 5427 hCoV-19/Thailand/Siriraj_5/2020|EPI_ISL_447908|202 .................... .................... .................... .................... .................... ....................
C 5428 hCoV-19/Thailand/Nonthaburi_59/2020|EPI_ISL_447909 .................... .................... .................... .................... .................... ....................
C 5429 hCoV-19/Thailand/Nonthaburi_68/2020|EPI_ISL_447910 .................... .................... .................... .................... .................... ....................
C 5430 hCoV-19/Belgium/UGent-67/2020|EPI_ISL_451178|2020- .................... .................... .................... .................... .................... ....................
C 5431 hCoV-19/Belgium/UGent-68/2020|EPI_ISL_451179|2020- .................... .................... .................... .................... .................... ....................
C 5432 hCoV-19/Belgium/UGent-70/2020|EPI_ISL_451181|2020- .................... .................... .................... .................... .................... ....................
C 5433 hCoV-19/Uganda/UG001/2020|EPI_ISL_451183|2020-03-2 .................... .................... .................... .................... .................... ....................
C 5434 hCoV-19/Uganda/UG002/2020|EPI_ISL_451184|2020-03-2 .................... .................... .................... .................... .................... ....................
C 5435 hCoV-19/Uganda/UG004/2020|EPI_ISL_451186|2020-03-2 .................... .................... .................... .................... .................... ....................
C 5436 hCoV-19/Sweden/20-50292/2020|EPI_ISL_454900|2020-0 .................... .................... .................... .................... .................... ....................
C 5437 hCoV-19/Sweden/20-50293/2020|EPI_ISL_454901|2020-0 .................... .................... .................... .................... .................... ....................
C 5438 hCoV-19/Sweden/20-50294/2020|EPI_ISL_454902|2020-0 .................... .................... .................... .................... .................... ....................
C 5439 hCoV-19/Sweden/20-50268/2020|EPI_ISL_454903|2020-0 .................... .................... .................... .................... .................... ....................
C 5440 hCoV-19/Wuhan/HB-WH1-122/2020|EPI_ISL_454904|2020- .................... .................... .................... .................... .................... ....................
C 5441 hCoV-19/Wuhan/HB-WH1-124/2020|EPI_ISL_454905|2020- .................... .................... .................... .................... .................... ....................
C 5442 hCoV-19/Wuhan/HB-WH1-125/2020|EPI_ISL_454906|2020- .................... .................... .................... .................... .................... ....................
C 5443 hCoV-19/Wuhan/HB-WH1-126/2020|EPI_ISL_454907|2020- .................... .................... .................... .................... .................... ....................
C 5444 hCoV-19/Wuhan/HB-WH1-127/2020|EPI_ISL_454908|2020- .................... .................... .................... .................... .................... ....................
C 5445 hCoV-19/Wuhan/HB-WH1-130/2020|EPI_ISL_454909|2020- .................... .................... .................... .................... .................... ....................
C 5446 hCoV-19/Wuhan/HB-WH1-131/2020|EPI_ISL_454910|2020- .................... .................... .................... .................... .................... ....................
C 5447 hCoV-19/Wuhan/HB-WH1-134/2020|EPI_ISL_454911|2020- .................... .................... .................... .................... .................... ....................
C 5448 hCoV-19/Wuhan/HB-WH1-135/2020|EPI_ISL_454912|2020- .................... .................... .................... .................... .................... ....................
C 5449 hCoV-19/Wuhan/HB-WH1-136/2020|EPI_ISL_454913|2020- .................... .................... .................... .................... .................... ....................
C 5450 hCoV-19/Wuhan/HB-WH1-137/2020|EPI_ISL_454914|2020- .................... .................... .................... .................... .................... ....................
C 5451 hCoV-19/Wuhan/HB-WH1-138/2020|EPI_ISL_454915|2020- .................... .................... .................... .................... .................... ....................
C 5452 hCoV-19/Wuhan/HB-WH1-140/2020|EPI_ISL_454916|2020- .................... .................... .................... .................... .................... ....................
C 5453 hCoV-19/Wuhan/HB-WH1-141/2020|EPI_ISL_454917|2020- .................... .................... .................... .................... .................... ....................
C 5454 hCoV-19/Wuhan/HB-WH1-142/2020|EPI_ISL_454918|2020- .................... .................... .................... .................... .................... ....................
C 5455 hCoV-19/Wuhan/HB-WH1-143/2020|EPI_ISL_454919|2020- .................... .................... .................... .................... .................... ....................

| | | | | | | | |
|---|---|---|---|---|---|---|---|
| C | 5456 hCoV-19/Wuhan/HB-WH2-148/2020\|EPI_ISL_454920\|2020- | ···················· | ···················· | ···················· | ···················· | ···················· | ···················· |
| C | 5457 hCoV-19/Wuhan/HB-WH2-149/2020\|EPI_ISL_454921\|2020- | ···················· | ···················· | ···················· | ···················· | ··C················· | ···················· |
| C | 5458 hCoV-19/Wuhan/HB-WH2-155/2020\|EPI_ISL_454923\|2020- | ···················· | ···················· | ···················· | ···················· | ···················· | ···················· |
| C | 5459 hCoV-19/Wuhan/HB-WH2-156/2020\|EPI_ISL_454924\|2020- | ···················· | ···················· | ···················· | ···················· | ···················· | ···················· |
| C | 5460 hCoV-19/Wuhan/HB-WH2-158/2020\|EPI_ISL_454926\|2020- | ···················· | ·A·················· | ···················· | ···················· | ···················· | ···················· |
| C | 5461 hCoV-19/Wuhan/HB-WH2-159/2020\|EPI_ISL_454927\|2020- | ···················· | ···················· | ···················· | ···················· | ···················· | ···················· |
| C | 5462 hCoV-19/Wuhan/HB-WH2-160/2020\|EPI_ISL_454928\|2020- | ···················· | ···················· | ···················· | ···················· | ···················· | ···················· |
| C | 5463 hCoV-19/Wuhan/HB-WH2-161/2020\|EPI_ISL_454929\|2020- | ···················· | ···················· | ···················· | ···················· | ···················· | ···················· |
| C | 5464 hCoV-19/England/EXET-1357EE/2020\|EPI_ISL_457101\|20 | ···················· | ···················· | ···················· | ···················· | ···················· | ···················· |
| C | 5465 hCoV-19/England/EXET-135818/2020\|EPI_ISL_457103\|20 | ···················· | ···················· | ···················· | ···················· | ···················· | ···················· |
| C | 5466 hCoV-19/England/EXET-135827/2020\|EPI_ISL_457104\|20 | ···················· | ···················· | ···················· | ···················· | ···················· | ···················· |
| C | 5467 hCoV-19/England/EXET-135836/2020\|EPI_ISL_457105\|20 | ···················· | ···················· | ···················· | ···················· | ···················· | ···················· |
| C | 5468 hCoV-19/England/EXET-135845/2020\|EPI_ISL_457106\|20 | ···················· | ···················· | ···················· | ···················· | ···················· | ···················· |
| C | 5469 hCoV-19/England/EXET-135863/2020\|EPI_ISL_457108\|20 | ···················· | ···················· | ···················· | ···················· | ···················· | ···················· |
| C | 5470 hCoV-19/England/EXET-135872/2020\|EPI_ISL_457109\|20 | ···················· | ···················· | ···················· | ···················· | ···················· | ···················· |
| C | 5471 hCoV-19/England/EXET-1358AF/2020\|EPI_ISL_457111\|20 | ···················· | ···················· | ···················· | ···················· | ···················· | ···················· |
| C | 5472 hCoV-19/England/EXET-1358BE/2020\|EPI_ISL_457112\|20 | ···················· | ···················· | ···················· | ···················· | ···················· | ···················· |
| C | 5473 hCoV-19/England/EXET-1358EB/2020\|EPI_ISL_457114\|20 | ···················· | ···················· | ···················· | ···················· | ···················· | ···················· |
| C | 5474 hCoV-19/England/EXET-135906/2020\|EPI_ISL_457115\|20 | ···················· | ···················· | ···················· | ···················· | ···················· | ···················· |
| C | 5475 hCoV-19/England/EXET-135915/2020\|EPI_ISL_457116\|20 | ···················· | ···················· | ···················· | ···················· | ···················· | ···················· |
| C | 5476 hCoV-19/England/EXET-135933/2020\|EPI_ISL_457117\|20 | ···················· | ···················· | ···················· | ···················· | ···················· | ···················· |
| C | 5477 hCoV-19/England/EXET-13597F/2020\|EPI_ISL_457119\|20 | ···················· | ···················· | ···················· | ···················· | ···················· | ···················· |
| C | 5478 hCoV-19/England/EXET-1359CA/2020\|EPI_ISL_457122\|20 | ···················· | ···················· | ···················· | ···················· | ···················· | ···················· |
| C | 5479 hCoV-19/England/EXET-1359D9/2020\|EPI_ISL_457123\|20 | ···················· | ···················· | ···················· | ···················· | ···················· | ···················· |
| C | 5480 hCoV-19/England/EXET-1359F7/2020\|EPI_ISL_457124\|20 | ···················· | ···················· | ···················· | ···················· | ···················· | ···················· |
| C | 5481 hCoV-19/England/EXET-135A03/2020\|EPI_ISL_457125\|20 | ···················· | ···················· | ···················· | ···················· | ···················· | ···················· |
| C | 5482 hCoV-19/England/EXET-135A12/2020\|EPI_ISL_457126\|20 | ···················· | ···················· | ···················· | ···················· | ···················· | ···················· |
| C | 5483 hCoV-19/England/EXET-135A21/2020\|EPI_ISL_457127\|20 | ···················· | ···················· | ···················· | ···················· | ···················· | ···················· |
| C | 5484 hCoV-19/England/EXET-135A30/2020\|EPI_ISL_457128\|20 | ···················· | ···················· | ···················· | ···················· | ···················· | ···················· |
| C | 5485 hCoV-19/England/EXET-135A6D/2020\|EPI_ISL_457130\|20 | ···················· | ···················· | ···················· | ···················· | ···················· | ···················· |
| C | 5486 hCoV-19/England/EXET-135A8B/2020\|EPI_ISL_457131\|20 | ···················· | ···················· | ···················· | ···················· | ···················· | ···················· |
| C | 5487 hCoV-19/England/EXET-135B4C/2020\|EPI_ISL_457136\|20 | ···················· | ···················· | ···················· | ···················· | ···················· | ···················· |
| C | 5488 hCoV-19/England/EXET-135B5B/2020\|EPI_ISL_457137\|20 | ···················· | ···················· | ···················· | ···················· | ···················· | ···················· |
| C | 5489 hCoV-19/England/EXET-135DFB/2020\|EPI_ISL_457170\|20 | ···················· | ···················· | ···················· | ···················· | ···················· | ···················· |
| C | 5490 hCoV-19/England/EXET-135E34/2020\|EPI_ISL_457171\|20 | ···················· | ···················· | ···················· | ···················· | ···················· | ···················· |
| C | 5492 hCoV-19/England/EXET-135EAD/2020\|EPI_ISL_457176\|20 | ···················· | ···················· | ···················· | ···················· | ···················· | ···················· |
| C | 5493 hCoV-19/England/EXET-135EBC/2020\|EPI_ISL_457177\|20 | ···················· | ···················· | ···················· | ···················· | ···················· | ···················· |
| C | 5494 hCoV-19/England/EXET-135ECB/2020\|EPI_ISL_457178\|20 | ···················· | ···················· | ···················· | ···················· | ···················· | ···················· |
| C | 5495 hCoV-19/England/EXET-135EDA/2020\|EPI_ISL_457179\|20 | ···················· | ···················· | ···················· | ···················· | ···················· | ···················· |
| C | 5496 hCoV-19/England/EXET-135F04/2020\|EPI_ISL_457180\|20 | ···················· | ···················· | ···················· | ···················· | ···················· | ···················· |
| C | 5497 hCoV-19/England/EXET-135F13/2020\|EPI_ISL_457181\|20 | ···················· | ···················· | ···················· | ···················· | ···················· | ···················· |
| C | 5498 hCoV-19/England/EXET-135F31/2020\|EPI_ISL_457182\|20 | ···················· | ···················· | ···················· | ···················· | ···················· | ···················· |
| C | 5499 hCoV-19/England/EXET-135F7D/2020\|EPI_ISL_457184\|20 | ···················· | ···················· | ···················· | ···················· | ···················· | ···················· |
| C | 5500 hCoV-19/England/EXET-135F8C/2020\|EPI_ISL_457185\|20 | ···················· | ···················· | ···················· | ···················· | ···················· | ···················· |
| C | 5501 hCoV-19/England/EXET-136000/2020\|EPI_ISL_457189\|20 | ···················· | ···················· | ···················· | ···················· | ···················· | ···················· |
| C | 5503 hCoV-19/England/EXET-13606A/2020\|EPI_ISL_457192\|20 | ···················· | ···················· | ···················· | ···················· | ···················· | ···················· |
| C | 5505 hCoV-19/England/EXET-1360B5/2020\|EPI_ISL_457195\|20 | ···················· | ···················· | ···················· | ···················· | ···················· | ···················· |
| C | 5507 hCoV-19/England/EXET-1360F1/2020\|EPI_ISL_457198\|20 | ···················· | ···················· | ···················· | ···················· | ···················· | ···················· |
| C | 5508 hCoV-19/England/BRIS-12AF7B/2020\|EPI_ISL_458400\|20 | ···················· | ···················· | ···················· | ···················· | ···················· | ···················· |
| C | 5509 hCoV-19/England/BRIS-12AB1D/2020\|EPI_ISL_458403\|20 | ···················· | ···················· | ···················· | ···················· | ···················· | ···················· |
| C | 5510 hCoV-19/England/BRIS-129FC7/2020\|EPI_ISL_458405\|20 | ···················· | ···················· | ···················· | ···················· | ···················· | ···················· |
| C | 5511 hCoV-19/England/BRIS-12A773/2020\|EPI_ISL_458406\|20 | ···················· | ···················· | ···················· | ···················· | ···················· | ···················· |
| C | 5512 hCoV-19/England/BRIS-129DCD/2020\|EPI_ISL_458407\|20 | ···················· | ···················· | ···················· | ···················· | ···················· | ···················· |
| C | 5513 hCoV-19/England/BRIS-12A5A6/2020\|EPI_ISL_458410\|20 | ···················· | ···················· | ···················· | ···················· | ···················· | ···················· |
| C | 5514 hCoV-19/England/BRIS-12A12A/2020\|EPI_ISL_458412\|20 | ···················· | ···················· | ···················· | ···················· | ···················· | ···················· |
| C | 5515 hCoV-19/England/BRIS-12A38E/2020\|EPI_ISL_458413\|20 | ···················· | ···················· | ···················· | ···················· | ···················· | ···················· |
| C | 5516 hCoV-19/England/BRIS-12A870/2020\|EPI_ISL_458414\|20 | ···················· | ···················· | ···················· | ···················· | ···················· | ···················· |
| C | 5517 hCoV-19/England/BRIS-12A2CD/2020\|EPI_ISL_458415\|20 | ···················· | ···················· | ···················· | ···················· | ···················· | ···················· |
| C | 5518 hCoV-19/England/BRIS-129C48/2020\|EPI_ISL_458416\|20 | ···················· | ···················· | ···················· | ···················· | ···················· | ···················· |
| C | 5519 hCoV-19/England/BRIS-129CA2/2020\|EPI_ISL_458419\|20 | ···················· | ···················· | ···················· | ···················· | ···················· | ···················· |
| C | 5520 hCoV-19/England/BRIS-12A9F5/2020\|EPI_ISL_458420\|20 | ···················· | ···················· | ···················· | ···················· | ···················· | ···················· |
| C | 5521 hCoV-19/England/BRIS-12A351/2020\|EPI_ISL_458421\|20 | ···················· | ···················· | ···················· | ···················· | ···················· | ···················· |
| C | 5522 hCoV-19/England/BRIS-12A1DF/2020\|EPI_ISL_458422\|20 | ···················· | ···················· | ···················· | ···················· | ···················· | ···················· |
| C | 5523 hCoV-19/England/BRIS-12A3BB/2020\|EPI_ISL_458423\|20 | ···················· | ···················· | ···················· | ···················· | ···················· | ···················· |
| C | 5524 hCoV-19/England/BRIS-129844/2020\|EPI_ISL_458425\|20 | ···················· | ···················· | ···················· | ···················· | ···················· | ···················· |
| C | 5525 hCoV-19/England/BRIS-1298AE/2020\|EPI_ISL_458426\|20 | ···················· | ···················· | ···················· | ···················· | ···················· | ···················· |
| C | 5526 hCoV-19/England/BRIS-12ADBD/2020\|EPI_ISL_458427\|20 | ···················· | ···················· | ···················· | ···················· | ···················· | ···················· |
| C | 5527 hCoV-19/England/BRIS-129A20/2020\|EPI_ISL_458430\|20 | ···················· | ···················· | ···················· | ···················· | ···················· | ···················· |
| C | 5528 hCoV-19/England/BRIS-12A755/2020\|EPI_ISL_458431\|20 | ···················· | ···················· | ···················· | ···················· | ···················· | ···················· |
| C | 5529 hCoV-19/England/BRIS-12AC38/2020\|EPI_ISL_458434\|20 | ···················· | ···················· | ···················· | ···················· | ···················· | ···················· |
| C | 5530 hCoV-19/England/BRIS-12A02D/2020\|EPI_ISL_458435\|20 | ···················· | ···················· | ···················· | ···················· | ···················· | ···················· |
| C | 5531 hCoV-19/England/BRIS-1298F9/2020\|EPI_ISL_458436\|20 | ···················· | ···················· | ···················· | ···················· | ···················· | ···················· |
| C | 5532 hCoV-19/England/BRIS-12AD26/2020\|EPI_ISL_458437\|20 | ···················· | ···················· | ···················· | ···················· | ···················· | ···················· |
| C | 5533 hCoV-19/England/BRIS-12A4D6/2020\|EPI_ISL_458438\|20 | ···················· | ···················· | ···················· | ···················· | ···················· | ···················· |
| C | 5534 hCoV-19/England/BRIS-12A175/2020\|EPI_ISL_458439\|20 | ···················· | ···················· | ···················· | ···················· | ···················· | ···················· |
| C | 5535 hCoV-19/England/BRIS-129905/2020\|EPI_ISL_458440\|20 | ···················· | ···················· | ···················· | ···················· | ···················· | ···················· |
| C | 5536 hCoV-19/England/BRIS-12AC47/2020\|EPI_ISL_458441\|20 | ···················· | ···················· | ···················· | ···················· | ···················· | ···················· |
| C | 5537 hCoV-19/England/BRIS-12AB59/2020\|EPI_ISL_458442\|20 | ···················· | ···················· | ···················· | ···················· | ···················· | ···················· |

| | | |
|---|---|---|
| C | 5538 hCoV-19/England/BRIS-12A139/2020\|EPI_ISL_458443\|20 | ···················· ···················· ···················· ···················· ···················· ···················· |
| C | 5539 hCoV-19/England/BRIS-12A60D/2020\|EPI_ISL_458445\|20 | ···················· ···················· ···················· ···················· ···················· ···················· |
| C | 5540 hCoV-19/England/BRIS-12A99B/2020\|EPI_ISL_458446\|20 | ···················· ···················· ···················· ···················· ···················· ···················· |
| C | 5541 hCoV-19/England/BRIS-12ADAE/2020\|EPI_ISL_458447\|20 | ···················· ···················· ···················· ···················· ···················· ···················· |
| C | 5542 hCoV-19/England/BRIS-1298CC/2020\|EPI_ISL_458448\|20 | ···················· ···················· ···················· ···················· ···················· ···················· |
| C | 5543 hCoV-19/England/BRIS-129FD6/2020\|EPI_ISL_458449\|20 | ···················· ···················· ···················· ···················· ···················· ···················· |
| C | 5544 hCoV-19/England/BRIS-12A04B/2020\|EPI_ISL_458451\|20 | ···················· ···················· ···················· ···················· ···················· ···················· |
| C | 5545 hCoV-19/England/BRIS-12A3E8/2020\|EPI_ISL_458452\|20 | ···················· ···················· ···················· ···················· ···················· ···················· |
| C | 5546 hCoV-19/England/BRIS-12ABD1/2020\|EPI_ISL_458453\|20 | ···················· ···················· ···················· ···················· ···················· ···················· |
| C | 5547 hCoV-19/England/BRIS-12989F/2020\|EPI_ISL_458455\|20 | ···················· ···················· ···················· ···················· ···················· ···················· |
| C | 5548 hCoV-19/England/BRIS-12A209/2020\|EPI_ISL_458456\|20 | ···················· ···················· ···················· ···················· ···················· ···················· |
| C | 5549 hCoV-19/England/BRIS-12A1B1/2020\|EPI_ISL_458457\|20 | ···················· ···················· ···················· ···················· ···················· ···················· |
| C | 5550 hCoV-19/England/BRIS-129D63/2020\|EPI_ISL_458459\|20 | ···················· ···················· ···················· ···················· ···················· ···················· |
| C | 5551 hCoV-19/England/BRIS-12A825/2020\|EPI_ISL_458460\|20 | ···················· ···················· ···················· ···················· ···················· ···················· |
| C | 5552 hCoV-19/England/BRIS-12A7FB/2020\|EPI_ISL_458461\|20 | ···················· ···················· ···················· ···················· ···················· ···················· |
| C | 5553 hCoV-19/England/BRIS-12AAF2/2020\|EPI_ISL_458462\|20 | ···················· ···················· ···················· ···················· ···················· ···················· |
| C | 5554 hCoV-19/Scotland/GCVR-170CAC/2020\|EPI_ISL_459703\|2 | ···················· ···················· ···················· ···················· ···················· ···················· |
| C | 5555 hCoV-19/Scotland/GCVR-170CF7/2020\|EPI_ISL_459704\|2 | ···················· ·A·················· ···················· ···················· ···················· ···················· |
| C | 5556 hCoV-19/Scotland/GCVR-17093C/2020\|EPI_ISL_459705\|2 | ···················· ···················· ···················· ···················· ···················· ···················· |
| C | 5557 hCoV-19/Scotland/GCVR-16FF5C/2020\|EPI_ISL_459707\|2 | ···················· ···················· ···················· ···················· ···················· ···················· |
| C | 5558 hCoV-19/Scotland/GCVR-17125E/2020\|EPI_ISL_459708\|2 | ···················· ···················· ···················· ···················· ···················· ···················· |
| C | 5559 hCoV-19/Scotland/GCVR-170468/2020\|EPI_ISL_459709\|2 | ···················· ···················· ···················· ···················· ···················· ···················· |
| C | 5560 hCoV-19/Scotland/GCVR-1705B0/2020\|EPI_ISL_459710\|2 | ···················· ···················· ···················· ···················· ···················· ···················· |
| C | 5561 hCoV-19/Scotland/GCVR-1703D4/2020\|EPI_ISL_459711\|2 | ···················· ···················· ···················· ···················· ···················· ···················· |
| C | 5562 hCoV-19/Scotland/GCVR-170547/2020\|EPI_ISL_459712\|2 | ···················· ·A·················· ···················· ···················· ···················· ···················· |
| C | 5563 hCoV-19/Scotland/GCVR-170899/2020\|EPI_ISL_459713\|2 | ···················· ···················· ···················· ···················· ···················· ···················· |
| C | 5564 hCoV-19/Scotland/GCVR-170AA2/2020\|EPI_ISL_459714\|2 | ···················· ···················· ···················· ···················· ···················· ···················· |
| C | 5565 hCoV-19/Scotland/GCVR-17136A/2020\|EPI_ISL_459716\|2 | ···················· ···················· ···················· ···················· ···················· ···················· |
| C | 5566 hCoV-19/Scotland/GCVR-170C24/2020\|EPI_ISL_459718\|2 | ···················· ···················· ···················· ···················· ···················· ···················· |
| C | 5567 hCoV-19/Scotland/GCVR-17037A/2020\|EPI_ISL_459720\|2 | ···················· ···················· ···················· ···················· ···················· ···················· |
| C | 5568 hCoV-19/Scotland/GCVR-170741/2020\|EPI_ISL_459721\|2 | ···················· ···················· ···················· ···················· ···················· ···················· |
| C | 5569 hCoV-19/Scotland/GCVR-16FFA7/2020\|EPI_ISL_459724\|2 | ···················· ···················· ···················· ···················· ···················· ···················· |
| C | 5570 hCoV-19/England/BRIS-123868/2020\|EPI_ISL_459725\|20 | ···················· ···················· ···················· ···················· ···················· ···················· |
| C | 5571 hCoV-19/England/BRIS-12384A/2020\|EPI_ISL_459726\|20 | ···················· ····A··············· ···················· ···················· ···················· ···················· |
| C | 5572 hCoV-19/Luxembourg/LNS6387933/2020\|EPI_ISL_459900\| | ···················· ···················· ···················· ···················· ···················· ···················· |
| C | 5573 hCoV-19/Netherlands/NA_527/2020\|EPI_ISL_460700\|202 | ···················· ···················· ···················· ···················· ···················· ···················· |
| C | 5574 hCoV-19/Netherlands/NA_528/2020\|EPI_ISL_460701\|202 | ···················· ···················· ···················· ···················· ···················· ···················· |
| C | 5575 hCoV-19/Netherlands/NA_529/2020\|EPI_ISL_460702\|202 | ···················· ···················· ···················· ···················· ···················· ···················· |
| C | 5576 hCoV-19/Netherlands/NA_530/2020\|EPI_ISL_460703\|202 | ···················· ···················· ···················· ···················· ···················· ···················· |
| C | 5577 hCoV-19/Netherlands/NA_531/2020\|EPI_ISL_460704\|202 | ···················· ···················· ···················· ···················· ···················· ···················· |
| C | 5578 hCoV-19/Netherlands/NA_532/2020\|EPI_ISL_460705\|202 | ···················· ···················· ···················· ···················· ···················· ···················· |
| C | 5579 hCoV-19/Netherlands/NA_533/2020\|EPI_ISL_460706\|202 | ···················· ···················· ···················· ···················· ···················· ···················· |
| C | 5580 hCoV-19/Netherlands/NA_534/2020\|EPI_ISL_460707\|202 | ···················· ···················· ···················· ···················· ···················· ···················· |
| C | 5581 hCoV-19/Netherlands/NA_535/2020\|EPI_ISL_460708\|202 | ···················· ···················· ···················· ···················· ···················· ···················· |
| C | 5582 hCoV-19/Netherlands/NA_536/2020\|EPI_ISL_460709\|202 | ···················· ···················· ···················· ···················· ···················· ···················· |
| C | 5583 hCoV-19/Netherlands/NA_537/2020\|EPI_ISL_460710\|202 | ···················· ···················· ···················· ···················· ···················· ···················· |
| C | 5584 hCoV-19/Netherlands/NA_538/2020\|EPI_ISL_460711\|202 | ···················· ···················· ···················· ···················· ···················· ···················· |
| C | 5585 hCoV-19/Netherlands/NA_539/2020\|EPI_ISL_460712\|202 | ···················· ···················· ···················· ···················· ···················· ···················· |
| C | 5586 hCoV-19/Netherlands/NA_540/2020\|EPI_ISL_460713\|202 | ···················· ···················· ···················· ···················· ···················· ···················· |
| C | 5587 hCoV-19/Netherlands/NA_543/2020\|EPI_ISL_460716\|202 | ···················· ···················· ···················· ···················· ···················· ···················· |
| C | 5588 hCoV-19/Netherlands/NA_547/2020\|EPI_ISL_460717\|202 | ···················· ···················· ···················· ···················· ···················· ···················· |
| C | 5589 hCoV-19/Netherlands/NA_549/2020\|EPI_ISL_460718\|202 | ···················· ···················· ···················· ···················· ···················· ···················· |
| C | 5590 hCoV-19/Netherlands/NA_552/2020\|EPI_ISL_460720\|202 | ···················· ···················· ···················· ···················· ···················· ···················· |
| C | 5591 hCoV-19/Netherlands/NA_556/2020\|EPI_ISL_460722\|202 | ···················· ···········A········ ···················· ···················· ···················· ···················· |
| C | 5592 hCoV-19/Netherlands/NA_558/2020\|EPI_ISL_460724\|202 | ···················· ···················· ···················· ···················· ···················· ···················· |
| C | 5593 hCoV-19/Netherlands/NA_560/2020\|EPI_ISL_460725\|202 | ···················· ···················· ···················· ···················· ···················· ···················· |
| C | 5594 hCoV-19/Netherlands/NA_561/2020\|EPI_ISL_460726\|202 | ···················· ···················· ···················· ···················· ···················· ···················· |
| C | 5595 hCoV-19/Netherlands/NA_562/2020\|EPI_ISL_460727\|202 | ···················· ···················· ···················· ···················· ···················· ···················· |
| C | 5596 hCoV-19/Netherlands/NA_563/2020\|EPI_ISL_460728\|202 | ···················· ···················· ···················· ···················· ···················· ···················· |
| C | 5597 hCoV-19/Netherlands/NA_564/2020\|EPI_ISL_460729\|202 | ···················· ···················· ···················· ···················· ···················· ···················· |
| C | 5598 hCoV-19/Netherlands/NA_566/2020\|EPI_ISL_460730\|202 | ···················· ···················· ···················· ···················· ···················· ···················· |
| C | 5599 hCoV-19/Netherlands/NA_567/2020\|EPI_ISL_460731\|202 | ···················· ···················· ···················· ···················· ···················· ···················· |
| C | 5600 hCoV-19/Netherlands/NA_568/2020\|EPI_ISL_460732\|202 | ···················· ···················· ···················· ···················· ···················· ···················· |
| C | 5601 hCoV-19/Netherlands/NA_569/2020\|EPI_ISL_460733\|202 | ···················· ···················· ···················· ···················· ···················· ···················· |
| C | 5602 hCoV-19/Netherlands/NA_570/2020\|EPI_ISL_460734\|202 | ···················· ···················· ···················· ···················· ···················· ···················· |
| C | 5603 hCoV-19/Netherlands/NA_571/2020\|EPI_ISL_460735\|202 | ···················· ···················· ···················· ···················· ···················· ···················· |
| C | 5604 hCoV-19/Netherlands/NA_572/2020\|EPI_ISL_460736\|202 | ···················· ···················· ···················· ···················· ···················· ···················· |
| C | 5605 hCoV-19/Netherlands/NA_573/2020\|EPI_ISL_460737\|202 | ···················· ···················· ···················· ···················· ···················· ···················· |
| C | 5606 hCoV-19/Netherlands/NA_575/2020\|EPI_ISL_460739\|202 | ···················· ···················· ···················· ···················· ···················· ···················· |
| C | 5607 hCoV-19/Netherlands/NA_576/2020\|EPI_ISL_460740\|202 | ···················· ···················· ···················· ···················· ···················· ···················· |
| C | 5608 hCoV-19/Netherlands/NA_577/2020\|EPI_ISL_460741\|202 | ···················· ···················· ···················· ···················· ···················· ···················· |
| C | 5609 hCoV-19/Netherlands/NA_578/2020\|EPI_ISL_460742\|202 | ···················· ···················· ···················· ···················· ···················· ···················· |
| C | 5610 hCoV-19/Netherlands/NA_579/2020\|EPI_ISL_460743\|202 | ···················· ···················· ···················· ···················· ···················· ···················· |
| C | 5611 hCoV-19/Netherlands/NA_582/2020\|EPI_ISL_460746\|202 | ···················· ···················· ···················· ···················· ···················· ···················· |
| C | 5612 hCoV-19/Netherlands/NA_583/2020\|EPI_ISL_460747\|202 | ···················· ···················· ···················· ···················· ···················· ···················· |
| C | 5613 hCoV-19/Netherlands/NA_584/2020\|EPI_ISL_460748\|202 | ···················· ···················· ···················· ···················· ···················· ···················· |
| C | 5614 hCoV-19/Netherlands/NA_585/2020\|EPI_ISL_460749\|202 | ···················· ···················· ···················· ···················· ···················· ···················· |
| C | 5615 hCoV-19/Netherlands/NA_586/2020\|EPI_ISL_460750\|202 | ···················· ···················· ···················· ···················· ···················· ···················· |

| | | | | | | | | |
|---|---|---|---|---|---|---|---|---|
| C | 5616 | hCoV-19/Netherlands/NA_588/2020\|EPI_ISL_460752\|202 | ···················· | ···················· | ···················· | ···················· | ···················· | ···················· |
| C | 5617 | hCoV-19/Netherlands/NA_589/2020\|EPI_ISL_460753\|202 | ···················· | ···················· | ···················· | ···················· | ···················· | ···················· |
| C | 5618 | hCoV-19/Netherlands/NA_590/2020\|EPI_ISL_460754\|202 | ···················· | ···················· | ···················· | ···················· | ···················· | ···················· |
| C | 5619 | hCoV-19/Netherlands/NA_591/2020\|EPI_ISL_460755\|202 | ···················· | ···················· | ···················· | ···················· | ···················· | ···················· |
| C | 5620 | hCoV-19/Netherlands/NA_593/2020\|EPI_ISL_460756\|202 | ···················· | ···················· | ···················· | ···················· | ···················· | ···················· |
| C | 5621 | hCoV-19/Netherlands/NA_594/2020\|EPI_ISL_460757\|202 | ···················· | ···················· | ···················· | ···················· | ···················· | ···················· |
| C | 5622 | hCoV-19/Netherlands/NA_595/2020\|EPI_ISL_460758\|202 | ···················· | ···················· | ···················· | ···················· | ···················· | ···················· |
| C | 5623 | hCoV-19/Netherlands/NA_597/2020\|EPI_ISL_460760\|202 | ···················· | ···················· | ···················· | ···················· | ···················· | ···················· |
| C | 5624 | hCoV-19/Netherlands/NA_599/2020\|EPI_ISL_460762\|202 | ···················· | ···················· | ···················· | ···················· | ···················· | ···················· |
| C | 5625 | hCoV-19/Netherlands/NA_600/2020\|EPI_ISL_460763\|202 | ···················· | ···················· | ···················· | ···················· | ···················· | ···················· |
| C | 5626 | hCoV-19/Netherlands/NA_601/2020\|EPI_ISL_460764\|202 | ···················· | ···················· | ···················· | ···················· | ···················· | ···················· |
| C | 5627 | hCoV-19/Netherlands/NA_603/2020\|EPI_ISL_460766\|202 | ···················· | ···················· | ···················· | ···················· | ···················· | ···················· |
| C | 5628 | hCoV-19/Netherlands/NA_604/2020\|EPI_ISL_460767\|202 | ···················· | ···················· | ···················· | ···················· | ···················· | ···················· |
| C | 5629 | hCoV-19/Netherlands/NA_605/2020\|EPI_ISL_460768\|202 | ···················· | ···················· | ···················· | ···················· | ···················· | ···················· |
| C | 5630 | hCoV-19/Netherlands/NA_608/2020\|EPI_ISL_460771\|202 | ···················· | ···················· | ···················· | ···················· | ···················· | ···················· |
| C | 5631 | hCoV-19/Netherlands/NA_615/2020\|EPI_ISL_460778\|202 | ···················· | ···················· | ···················· | ···················· | ···················· | ···················· |
| C | 5632 | hCoV-19/Netherlands/NA_616/2020\|EPI_ISL_460779\|202 | ···················· | ···················· | ···················· | ···················· | ···················· | ···················· |
| C | 5633 | hCoV-19/USA/FL-UF10/2020\|EPI_ISL_447903\|2020-04-07 | ···················· | ···················· | ···················· | ···················· | ···················· | ···················· |
| C | 5634 | hCoV-19/Bangladesh/NIB-01/2020\|EPI_ISL_447904\|2020 | ···················· | ···················· | ···················· | ···················· | ···················· | ···················· |
| C | 5635 | hCoV-19/Thailand/Siriraj_1/2020\|EPI_ISL_447906\|202 | ···················· | ·A·················· | ···················· | ···················· | ···················· | ···················· |
| C | 5636 | hCoV-19/Thailand/Siriraj_2/2020\|EPI_ISL_447907\|202 | ···················· | ·A·················· | ···················· | ···················· | ···················· | ···················· |
| C | 5637 | hCoV-19/Thailand/Nonthaburi_193/2020\|EPI_ISL_44791 | ···················· | ···················· | ···················· | ···················· | ···················· | ···················· |
| C | 5638 | hCoV-19/Thailand/Bangkok_237/2020\|EPI_ISL_447912\|2 | ···················· | ···················· | ···················· | ···················· | ···················· | ···················· |
| C | 5639 | hCoV-19/Thailand/Bangkok_238/2020\|EPI_ISL_447913\|2 | ···················· | ···················· | ···················· | ···················· | ···················· | ···················· |
| C | 5640 | hCoV-19/Thailand/Phuket_247/2020\|EPI_ISL_447914\|20 | ···················· | ···················· | ···················· | ···················· | ···················· | ···················· |
| C | 5641 | hCoV-19/Thailand/Bangkok_269/2020\|EPI_ISL_447915\|2 | ···················· | ···················· | ···················· | ···················· | ···················· | ···················· |
| C | 5642 | hCoV-19/Thailand/Bangkok_323/2020\|EPI_ISL_447916\|2 | ···················· | ···················· | ···················· | ···················· | ···················· | ···················· |
| C | 5643 | hCoV-19/Thailand/Nonthaburi_363/2020\|EPI_ISL_44791 | ···················· | ···················· | ···················· | ···················· | ···················· | ···················· |
| C | 5644 | hCoV-19/Thailand/Bangkok_580/2020\|EPI_ISL_447918\|2 | ···················· | ···················· | ···················· | ···················· | ···················· | ···················· |
| C | 5645 | hCoV-19/Thailand/Samut prakarn_840/2020\|EPI_ISL_44 | ···················· | ···················· | ···················· | ···················· | ···················· | ···················· |
| C | 5646 | hCoV-19/Thailand/Bangkok_1716/2020\|EPI_ISL_447920\| | ···················· | ···················· | ···················· | ···················· | ···················· | ···················· |
| C | 5647 | hCoV-19/Thailand/Bangkok_2294/2020\|EPI_ISL_447921\| | ···················· | ···················· | ···················· | ···················· | ···················· | ···················· |
| C | 5648 | hCoV-19/England/BIRM-5E9DB/2020\|EPI_ISL_447927\|202 | ···················· | ···················· | ···················· | ···················· | ···················· | ···················· |
| C | 5649 | hCoV-19/England/BIRM-5E9EA/2020\|EPI_ISL_447928\|202 | ···················· | ···················· | ···················· | ···················· | ···················· | ···················· |
| C | 5653 | hCoV-19/England/BIRM-5ECE1/2020\|EPI_ISL_447939\|202 | ···················· | ···················· | ···················· | ···················· | ···················· | ···················· |
| C | 5655 | hCoV-19/England/CAMB-1ACFDB/2020\|EPI_ISL_447953\|20 | ···················· | ···················· | ···················· | ···················· | ···················· | ···················· |
| C | 5656 | hCoV-19/England/CAMB-1ACFEA/2020\|EPI_ISL_447954\|20 | ···················· | ···················· | ···················· | ···················· | ···················· | ···················· |
| C | 5657 | hCoV-19/England/CAMB-1ACFF9/2020\|EPI_ISL_447955\|20 | ···················· | ···A················ | ···················· | ···················· | ···················· | ···················· |
| C | 5658 | hCoV-19/England/CAMB-1AD004/2020\|EPI_ISL_447956\|20 | ···················· | ···················· | ···················· | ···················· | ···················· | ···················· |
| C | 5659 | hCoV-19/England/CAMB-1AD040/2020\|EPI_ISL_447959\|20 | ···················· | ···················· | ···················· | ···················· | ···················· | ···················· |
| C | 5660 | hCoV-19/England/CAMB-1AD06E/2020\|EPI_ISL_447960\|20 | ···················· | ···················· | ···················· | ···················· | ···················· | ···················· |
| C | 5661 | hCoV-19/England/CAMB-1AD0AA/2020\|EPI_ISL_447963\|20 | ···················· | ···················· | ···················· | ···················· | ···················· | ···················· |
| C | 5662 | hCoV-19/England/CAMB-1AD0B9/2020\|EPI_ISL_447964\|20 | ···················· | ···················· | ···················· | ···················· | ···················· | ···················· |
| C | 5663 | hCoV-19/Australia/SAP061/2020\|EPI_ISL_451100\|2020- | ···················· | ···················· | ···················· | ···················· | ···················· | ···················· |
| C | 5664 | hCoV-19/Australia/SAP062/2020\|EPI_ISL_451101\|2020- | ···················· | ···················· | ···················· | ···················· | ···················· | ···················· |
| C | 5665 | hCoV-19/Australia/SAP072/2020\|EPI_ISL_451109\|2020- | ···················· | ···················· | ···················· | ···················· | ···················· | ···················· |
| C | 5666 | hCoV-19/Australia/SAP075/2020\|EPI_ISL_451112\|2020- | ···················· | ···················· | ···················· | ···················· | ···················· | ···················· |
| C | 5667 | hCoV-19/Australia/SAP076/2020\|EPI_ISL_451113\|2020- | ···················· | ···················· | ···················· | ···················· | ···················· | ···················· |
| C | 5668 | hCoV-19/Australia/SAP077/2020\|EPI_ISL_451114\|2020- | ···················· | ···················· | ···················· | ···················· | ···················· | ···················· |
| C | 5669 | hCoV-19/Australia/SAP079/2020\|EPI_ISL_451115\|2020- | ···················· | ···················· | ···················· | ···················· | ···················· | ···················· |
| C | 5670 | hCoV-19/Australia/SAP080/2020\|EPI_ISL_451116\|2020- | ···················· | ···················· | ···················· | ···················· | ···················· | ···················· |
| C | 5671 | hCoV-19/Australia/SAP081/2020\|EPI_ISL_451117\|2020- | ···················· | ···················· | ···················· | ···················· | ···················· | ···················· |
| C | 5672 | hCoV-19/Australia/SAP082/2020\|EPI_ISL_451118\|2020- | ···················· | ···················· | ···················· | ···················· | ···················· | ···················· |
| C | 5673 | hCoV-19/Australia/SAP085/2020\|EPI_ISL_451121\|2020- | ···················· | ···················· | ···················· | ···················· | ···················· | ···················· |
| C | 5674 | hCoV-19/Australia/SAP087/2020\|EPI_ISL_451122\|2020- | ···················· | ···················· | ···················· | ···················· | ···················· | ···················· |
| C | 5675 | hCoV-19/Australia/SAP088/2020\|EPI_ISL_451123\|2020- | ···················· | ···················· | ···················· | ···················· | ···················· | ···················· |
| C | 5676 | hCoV-19/Australia/SAP090/2020\|EPI_ISL_451125\|2020- | ···················· | ···················· | ···················· | ···················· | ···················· | ···················· |
| C | 5677 | hCoV-19/Australia/SAP091/2020\|EPI_ISL_451126\|2020- | ···················· | ···················· | ···················· | ···················· | ···················· | ···················· |
| C | 5678 | hCoV-19/Australia/SAP093/2020\|EPI_ISL_451128\|2020- | ···················· | ···················· | ···················· | ···················· | ···················· | ···················· |
| C | 5679 | hCoV-19/Australia/SAP095/2020\|EPI_ISL_451129\|2020- | ···················· | ···················· | ···················· | ···················· | ···················· | ···················· |
| C | 5680 | hCoV-19/Australia/SAP111/2020\|EPI_ISL_451131\|2020- | ···················· | ···················· | ···················· | ···················· | ···················· | ···················· |
| C | 5681 | hCoV-19/Australia/SAP115/2020\|EPI_ISL_451132\|2020- | ···················· | ···················· | ···················· | ···················· | ···················· | ···················· |
| C | 5682 | hCoV-19/Australia/SAP122/2020\|EPI_ISL_451133\|2020- | ···················· | ···················· | ···················· | ···················· | ···················· | ···················· |
| C | 5683 | hCoV-19/Australia/SAP125/2020\|EPI_ISL_451134\|2020- | ···················· | ···················· | ···················· | ···················· | ···················· | ···················· |
| C | 5684 | hCoV-19/Australia/SAP126/2020\|EPI_ISL_451135\|2020- | ···················· | ···················· | ···················· | ···················· | ···················· | ···················· |
| C | 5685 | hCoV-19/Australia/SAP127/2020\|EPI_ISL_451136\|2020- | ···················· | ···················· | ···················· | ···················· | ···················· | ···················· |
| C | 5686 | hCoV-19/Australia/SAP130/2020\|EPI_ISL_451137\|2020- | ···················· | ···················· | ···················· | ···················· | ···················· | ···················· |
| C | 5687 | hCoV-19/Australia/SAP133/2020\|EPI_ISL_451138\|2020- | ···················· | ···················· | ···················· | ···················· | ···················· | ···················· |
| C | 5688 | hCoV-19/Australia/SAP134/2020\|EPI_ISL_451139\|2020- | ···················· | ···················· | ···················· | ···················· | ···················· | ···················· |
| C | 5689 | hCoV-19/Australia/SAP136/2020\|EPI_ISL_451140\|2020- | ···················· | ···················· | ···················· | ···················· | ···················· | ···················· |
| C | 5690 | hCoV-19/Australia/SAP137/2020\|EPI_ISL_451141\|2020- | ···················· | ···················· | ···················· | ···················· | ···················· | ···················· |
| C | 5691 | hCoV-19/Australia/SAP146/2020\|EPI_ISL_451144\|2020- | ···················· | ···················· | ···················· | ···················· | ···················· | ···················· |
| C | 5692 | hCoV-19/Australia/SAP147/2020\|EPI_ISL_451145\|2020- | ···················· | ···················· | ···················· | ···················· | ···················· | ···················· |
| C | 5693 | hCoV-19/Australia/SAP153/2020\|EPI_ISL_451147\|2020- | ···················· | ···················· | ···················· | ···················· | ···················· | ···················· |
| C | 5694 | hCoV-19/Australia/SAP154/2020\|EPI_ISL_451148\|2020- | ···················· | ···················· | ···················· | ···················· | ···················· | ···················· |
| C | 5695 | hCoV-19/India/GJ-GBRC104/2020\|EPI_ISL_451149\|2020- | ···················· | ·A·················· | ···················· | ···················· | ···················· | ···A················ |
| C | 5696 | hCoV-19/India/GJ-GBRC105/2020\|EPI_ISL_451150\|2020- | ···················· | ···················· | ···················· | ···················· | ···················· | ···················· |
| C | 5697 | hCoV-19/India/GJ-GBRC106/2020\|EPI_ISL_451151\|2020- | ···················· | ·A·················· | ···················· | ···················· | ···················· | ···A················ |

| | | | | | | | | |
|---|---|---|---|---|---|---|---|---|
| C | 5698 hCoV-19/India/GJ-GBRC107/2020\|EPI_ISL_451152\|2020- | ···················· | ·A·················· | ···················· | ···················· | ···················· | ··A················· | |
| C | 5699 hCoV-19/India/GJ-GBRC108/2020\|EPI_ISL_451153\|2020- | ···················· | ·A·················· | ···················· | ···················· | ···················· | ··A················· | |
| C | 5700 hCoV-19/India/GJ-GBRC109/2020\|EPI_ISL_451154\|2020- | ···················· | ···················· | ···················· | ···················· | ···················· | ···················· | |
| C | 5701 hCoV-19/India/GJ-GBRC110/2020\|EPI_ISL_451155\|2020- | ···················· | ···················· | ···················· | ···················· | ···················· | ···················· | |
| C | 5702 hCoV-19/India/GJ-GBRC111/2020\|EPI_ISL_451156\|2020- | ···················· | ···················· | ···················· | ···················· | ···················· | ···················· | |
| C | 5703 hCoV-19/India/GJ-GBRC112/2020\|EPI_ISL_451157\|2020- | ···················· | ···················· | ···················· | ···················· | ···················· | ···················· | |
| C | 5704 hCoV-19/India/GJ-GBRC113/2020\|EPI_ISL_451158\|2020- | ···················· | ···················· | ···················· | ···················· | ···················· | ···················· | |
| C | 5705 hCoV-19/India/GJ-GBRC114/2020\|EPI_ISL_451159\|2020- | ···················· | ···················· | ···················· | ···················· | ···················· | ···················· | |
| C | 5706 hCoV-19/India/GJ-GBRC115/2020\|EPI_ISL_451160\|2020- | ···················· | ···················· | ···················· | ···················· | ···················· | ···················· | |
| C | 5707 hCoV-19/India/GJ-GBRC116/2020\|EPI_ISL_451161\|2020- | ···················· | ···················· | ···················· | ···················· | ···················· | ···················· | |
| C | 5708 hCoV-19/India/GJ-GBRC117/2020\|EPI_ISL_451162\|2020- | ···················· | ·A·················· | ···················· | ···················· | ···················· | ··A················· | |
| C | 5709 hCoV-19/India/GJ-GBRC118/2020\|EPI_ISL_451163\|2020- | ···················· | ···················· | ···················· | ···················· | ···················· | ···················· | |
| C | 5710 hCoV-19/Belgium/UGent-23/2020\|EPI_ISL_451164\|2020- | ···················· | ···················· | ···················· | ···················· | ···················· | ···················· | |
| C | 5711 hCoV-19/Belgium/UGent-53/2020\|EPI_ISL_451166\|2020- | ···················· | ···················· | ···················· | ···················· | ···················· | ···················· | |
| C | 5712 hCoV-19/Belgium/UGent-54/2020\|EPI_ISL_451167\|2020- | ···················· | ···················· | ···················· | ···················· | ···················· | ···················· | |
| C | 5713 hCoV-19/Belgium/UGent-57/2020\|EPI_ISL_451168\|2020- | ···················· | ···················· | ···················· | ···················· | ···················· | ···················· | |
| C | 5714 hCoV-19/Belgium/UGent-58/2020\|EPI_ISL_451169\|2020- | ···················· | ···················· | ···················· | ···················· | ···················· | ···················· | |
| C | 5715 hCoV-19/Belgium/UGent-59/2020\|EPI_ISL_451170\|2020- | ···················· | ···················· | ···················· | ···················· | ···················· | ···················· | |
| C | 5716 hCoV-19/Belgium/UGent-62/2020\|EPI_ISL_451173\|2020- | ···················· | ···················· | ···················· | ···················· | ···················· | ···················· | |
| C | 5717 hCoV-19/Belgium/UGent-64/2020\|EPI_ISL_451175\|2020- | ···················· | ···················· | ···················· | ···················· | ···················· | ···················· | |
| C | 5718 hCoV-19/Belgium/UGent-66/2020\|EPI_ISL_451177\|2020- | ···················· | ···················· | ···················· | ···················· | ···················· | ···················· | |
| C | 5719 hCoV-19/Belgium/UGent-69/2020\|EPI_ISL_451180\|2020- | ···················· | ···················· | ···················· | ···················· | ···················· | ···················· | |
| C | 5720 hCoV-19/Uganda/UG007/2020\|EPI_ISL_451189\|2020-03-3 | ···················· | ···················· | ···················· | ···················· | ···················· | ···················· | |
| C | 5721 hCoV-19/Uganda/UG008/2020\|EPI_ISL_451190\|2020-03-3 | ···················· | ···················· | ···················· | ···················· | ···················· | ·······NN··········· | |
| C | 5722 hCoV-19/Uganda/UG010/2020\|EPI_ISL_451192\|2020-03-3 | ···················· | ···················· | ···················· | ···················· | ···················· | ···················· | |
| C | 5723 hCoV-19/Uganda/UG011/2020\|EPI_ISL_451193\|2020-03-3 | ···················· | ···················· | ···················· | ···················· | ···················· | ···················· | |
| C | 5724 hCoV-19/Uganda/UG012/2020\|EPI_ISL_451194\|2020-03-2 | ···················· | ···················· | ···················· | ···················· | ···················· | ···················· | |
| C | 5725 hCoV-19/Uganda/UG013/2020\|EPI_ISL_451195\|2020-03-2 | ···················· | ···················· | ···················· | ···················· | ···················· | ···················· | |
| C | 5726 hCoV-19/Uganda/UG014/2020\|EPI_ISL_451196\|2020-03-2 | ···················· | ···················· | ···················· | ···················· | ···················· | ···················· | |
| C | 5727 hCoV-19/Uganda/UG015/2020\|EPI_ISL_451197\|2020-04-2 | ···················· | ···················· | ···················· | ···················· | ···················· | ···················· | |
| C | 5728 hCoV-19/Uganda/UG016/2020\|EPI_ISL_451198\|2020-04-2 | ···················· | ···················· | ···················· | ···················· | ···················· | ···················· | |
| C | 5729 hCoV-19/Uganda/UG017/2020\|EPI_ISL_451199\|2020-04-2 | ···················· | ···················· | ···················· | ···················· | ···················· | ···················· | |
| C | 5739 hCoV-19/Spain/AN-IBV-001969/2020\|EPI_ISL_452413\|20 | ···················· | ···················· | ···················· | ···················· | ··N················· | ·········N·········· | |
| C | 5746 hCoV-19/Spain/AN-IBV-001932/2020\|EPI_ISL_452443\|20 | ···················· | ···················· | ···················· | ···················· | ···················· | ···················· | |
| C | 5751 hCoV-19/Spain/AN-IBV-001922/2020\|EPI_ISL_452452\|20 | ···················· | ···················· | ···················· | ···················· | ···················· | ···················· | |
| C | 5774 hCoV-19/Spain/NC-IBV-001418/2020\|EPI_ISL_452483\|20 | ···················· | ···················· | ···················· | ···················· | ··N················· | ···················· | |
| C | 5776 hCoV-19/Spain/NC-IBV-001416/2020\|EPI_ISL_452485\|20 | ···················· | ···················· | ···················· | ···················· | ···················· | ···················· | |
| C | 5787 hCoV-19/England/SHEF-C7102/2020\|EPI_ISL_453721\|202 | ···················· | ···················· | ···················· | ···················· | ···················· | ···················· | |
| C | 5788 hCoV-19/England/SHEF-C724B/2020\|EPI_ISL_453722\|202 | ···················· | ···················· | ···················· | ···················· | ···················· | ···················· | |
| C | 5789 hCoV-19/England/SHEF-C7ECF/2020\|EPI_ISL_453724\|202 | ···················· | ···················· | ···················· | ···················· | ···················· | ···················· | |
| C | 5790 hCoV-19/England/SHEF-C822C/2020\|EPI_ISL_453728\|202 | ···················· | ···················· | ···················· | ···················· | ···················· | ···················· | |
| C | 5791 hCoV-19/England/SHEF-C88A2/2020\|EPI_ISL_453729\|202 | ···················· | ···················· | ···················· | ···················· | ···················· | ···················· | |
| C | 5792 hCoV-19/England/SHEF-C88DF/2020\|EPI_ISL_453730\|202 | ···················· | ···················· | ···················· | ···················· | ···················· | ···················· | |
| C | 5793 hCoV-19/England/SHEF-C8C4C/2020\|EPI_ISL_453732\|202 | ···················· | ···················· | ···················· | ···················· | ···················· | ···················· | |
| C | 5794 hCoV-19/England/SHEF-CA05D/2020\|EPI_ISL_453736\|202 | ···················· | ···················· | ···················· | ···················· | ···················· | ···················· | |
| C | 5795 hCoV-19/England/SHEF-CA4BB/2020\|EPI_ISL_453737\|202 | ···················· | ···················· | ···················· | ···················· | ···················· | ···················· | |
| C | 5796 hCoV-19/England/SHEF-CB159/2020\|EPI_ISL_453739\|202 | ···················· | ···················· | ···················· | ···················· | ···················· | ···················· | |
| C | 5797 hCoV-19/England/SHEF-CB292/2020\|EPI_ISL_453740\|202 | ···················· | ···················· | ···················· | ···················· | ···················· | ···················· | |
| C | 5798 hCoV-19/England/SHEF-CB423/2020\|EPI_ISL_453742\|202 | ···················· | ···················· | ···················· | ···················· | ···················· | ···················· | |
| C | 5799 hCoV-19/England/SHEF-CB502/2020\|EPI_ISL_453744\|202 | ···················· | ···················· | ···················· | ···················· | ···················· | ···················· | |
| C | 5800 hCoV-19/England/SHEF-CBA03/2020\|EPI_ISL_453745\|202 | ···················· | ···················· | ···················· | ···················· | ···················· | ···················· | |
| C | 5801 hCoV-19/England/SHEF-CC4AA/2020\|EPI_ISL_453746\|202 | ···················· | ···················· | ···················· | ···················· | ···················· | ···················· | |
| C | 5802 hCoV-19/England/SHEF-CCE7F/2020\|EPI_ISL_453748\|202 | ···················· | ···················· | ···················· | ···················· | ···················· | ···················· | |
| C | 5803 hCoV-19/England/SHEF-CD1DF/2020\|EPI_ISL_453749\|202 | ···················· | ···················· | ···················· | ···················· | ···················· | ···················· | |
| C | 5804 hCoV-19/England/SHEF-CD38E/2020\|EPI_ISL_453750\|202 | ···················· | ···················· | ···················· | ···················· | ···················· | ···················· | |
| C | 5805 hCoV-19/England/SHEF-CD500/2020\|EPI_ISL_453751\|202 | ···················· | ···················· | ···················· | ···················· | ···················· | ···················· | |
| C | 5806 hCoV-19/England/SHEF-CD807/2020\|EPI_ISL_453752\|202 | ···················· | ···················· | ···················· | ···················· | ···················· | ···················· | |
| C | 5807 hCoV-19/England/SHEF-CDA3E/2020\|EPI_ISL_453753\|202 | ···················· | ···················· | ···················· | ···················· | ···················· | ···················· | |
| C | 5808 hCoV-19/England/SHEF-CDD17/2020\|EPI_ISL_453755\|202 | ···················· | ···················· | ···················· | ···················· | ···················· | ···················· | |
| C | 5809 hCoV-19/England/SHEF-CDE8D/2020\|EPI_ISL_453756\|202 | ···················· | ···················· | ···················· | ···················· | ···················· | ···················· | |
| C | 5810 hCoV-19/England/SHEF-CF6ED/2020\|EPI_ISL_453758\|202 | ···················· | ···················· | ···················· | ···················· | ···················· | ···················· | |
| C | 5811 hCoV-19/England/SHEF-CF8BA/2020\|EPI_ISL_453759\|202 | ···················· | ···················· | ···················· | ···················· | ···················· | ···················· | |
| C | 5812 hCoV-19/England/SHEF-CFDAC/2020\|EPI_ISL_453760\|202 | ···················· | ···················· | ···················· | ···················· | ···················· | ···················· | |
| C | 5813 hCoV-19/England/SHEF-D01CA/2020\|EPI_ISL_453761\|202 | ···················· | ···················· | ···················· | ···················· | ···················· | ···················· | |
| C | 5814 hCoV-19/England/SHEF-D2CE5/2020\|EPI_ISL_453774\|202 | ···················· | ···················· | ···················· | ···················· | ···················· | ···················· | |
| C | 5815 hCoV-19/England/SHEF-D2DA6/2020\|EPI_ISL_453775\|202 | ···················· | ···················· | ···················· | ···················· | ···················· | ···················· | |
| C | 5816 hCoV-19/England/SHEF-D2E2B/2020\|EPI_ISL_453776\|202 | ···················· | ···················· | ···················· | ···················· | ···················· | ···················· | |
| C | 5817 hCoV-19/England/SHEF-D2F82/2020\|EPI_ISL_453777\|202 | ···················· | ···················· | ···················· | ···················· | ···················· | ···················· | |
| C | 5818 hCoV-19/England/NORT-2892CB/2020\|EPI_ISL_458600\|20 | ···················· | ···················· | ···················· | ···················· | ···················· | ···················· | |
| C | 5819 hCoV-19/England/NORT-2891A0/2020\|EPI_ISL_458601\|20 | ···················· | ···················· | ···················· | ···················· | ···················· | ···················· | |
| C | 5820 hCoV-19/England/NORT-288E5F/2020\|EPI_ISL_458602\|20 | ···················· | ···················· | ···················· | ···················· | ···················· | ···················· | |
| C | 5821 hCoV-19/England/NORT-288EAA/2020\|EPI_ISL_458603\|20 | ···················· | ···················· | ···················· | ···················· | ···················· | ···················· | |
| C | 5822 hCoV-19/England/NORT-28901C/2020\|EPI_ISL_458604\|20 | ···················· | ···················· | ···················· | ···················· | ···················· | ···················· | |
| C | 5823 hCoV-19/England/NORT-289744/2020\|EPI_ISL_458605\|20 | ···················· | ···················· | ···················· | ···················· | ···················· | ···················· | |
| C | 5824 hCoV-19/England/NORT-28944D/2020\|EPI_ISL_458606\|20 | ···················· | ···················· | ···················· | ···················· | ···················· | ···················· | |
| C | 5825 hCoV-19/England/NORT-28900D/2020\|EPI_ISL_458607\|20 | ···················· | ···················· | ···················· | ···················· | ···················· | ···················· | |
| C | 5826 hCoV-19/England/NORT-28947A/2020\|EPI_ISL_458609\|20 | ···················· | ···················A | ···················· | ···················· | ···················· | ···················· | 1 |
| C | 5827 hCoV-19/England/NORT-28997B/2020\|EPI_ISL_458610\|20 | ···················· | ···················· | ···················· | ···················· | ···················· | ···················· | |

| | | | | | | | |
|---|---|---|---|---|---|---|---|
| C | 5828 hCoV-19/England/NORT-289261/2020\|EPI_ISL_458611\|20 | ···················· | ···················· | ····················· | ···················· | ···················· | ···················· |
| C | 5829 hCoV-19/England/NORT-289410/2020\|EPI_ISL_458612\|20 | ···················· | ···················· | ····················· | ···················· | ···················· | ···················· |
| C | 5830 hCoV-19/England/NORT-2890B2/2020\|EPI_ISL_458613\|20 | ···················· | ···················· | ····················· | ···················· | ···················· | ···················· |
| C | 5831 hCoV-19/Luxembourg/LNS6469209/2020\|EPI_ISL_459901\| | ···················· | ···················· | ····················· | ···················· | ···················· | ···················· |
| C | 5832 hCoV-19/Luxembourg/LNS6862754/2020\|EPI_ISL_459903\| | ···················· | ···················· | ····················· | ···················· | ···················· | ···················· |
| C | 5833 hCoV-19/Luxembourg/LNS7342327/2020\|EPI_ISL_459905\| | ···················· | ···················· | ····················· | ···················· | ···················· | ···················· |
| C | 5834 hCoV-19/Luxembourg/LNS7820945/2020\|EPI_ISL_459906\| | ···················· | ···················· | ····················· | ···················· | ···················· | ···················· |
| C | 5835 hCoV-19/Harbin/HRB-26/2020\|EPI_ISL_459909\|2020-03- | ···················· | ···················· | ····················· | ···················· | ···················· | ···················· |
| C | 5836 hCoV-19/India/DL-MaxCov0003-CSIR-IGIB/2020\|EPI_ISL | ···················· | ···················· | ····················· | ···················· | ···················· | ···················· |
| C | 5837 hCoV-19/India/DL-MaxCov0010-CSIR-IGIB/2020\|EPI_ISL | ···················· | ···················· | ····················· | ···················· | ···················· | ···················· |
| C | 5838 hCoV-19/India/DL-MaxCov0012-CSIR-IGIB/2020\|EPI_ISL | ···················· | ···················· | ····················· | ···················· | ···················· | ···················· |
| C | 5839 hCoV-19/India/DL-MaxCov0013-CSIR-IGIB/2020\|EPI_ISL | ···················· | ···················· | ····················· | ···················· | ···················· | ···················· |
| C | 5840 hCoV-19/India/DL-MaxCov0014-CSIR-IGIB/2020\|EPI_ISL | ···················· | ···················· | ····················· | ···················· | ···················· | ···················· |
| C | 5841 hCoV-19/India/DL-MaxCov0015-CSIR-IGIB/2020\|EPI_ISL | ···················· | ···················· | ····················· | ···················· | ···················· | ···················· |
| C | 5842 hCoV-19/India/DL-MaxCov0016-CSIR-IGIB/2020\|EPI_ISL | ···················· | ···················· | ····················· | ···················· | ···················· | ···················· |
| C | 5843 hCoV-19/India/DL-MaxCov0017-CSIR-IGIB/2020\|EPI_ISL | ···················· | ···················· | ····················· | ···················· | ···················· | ···················· |
| C | 5844 hCoV-19/India/DL-MaxCov0018-CSIR-IGIB/2020\|EPI_ISL | ···················· | ···················· | ····················· | ···················· | ···················· | ···················· |
| C | 5845 hCoV-19/India/DL-MaxCov0019-CSIR-IGIB/2020\|EPI_ISL | ···················· | ···················· | ····················· | ···················· | ···················· | ···················· |
| C | 5846 hCoV-19/India/DL-MaxCov0020-CSIR-IGIB/2020\|EPI_ISL | ···················· | ···················· | ····················· | ···················· | ···················· | ···················· |
| C | 5847 hCoV-19/India/DL-MaxCov0021-CSIR-IGIB/2020\|EPI_ISL | ···················· | ···················· | ····················· | ···················· | ···················· | ···················· |
| C | 5848 hCoV-19/India/DL-MaxCov0022-CSIR-IGIB/2020\|EPI_ISL | ···················· | ···················· | ····················· | ···················· | ···················· | ···················· |
| C | 5849 hCoV-19/India/DL-MaxCov0024-CSIR-IGIB/2020\|EPI_ISL | ···················· | ···················· | ····················· | ···················· | ···················· | ···················· |
| C | 5850 hCoV-19/India/DL-MaxCov0025-CSIR-IGIB/2020\|EPI_ISL | ···················· | ···················· | ····················· | ···················· | ···················· | ···················· |
| C | 5851 hCoV-19/India/DL-MaxCov0030-CSIR-IGIB/2020\|EPI_ISL | ···················· | ···················· | ····················· | ···················· | ···················· | ···················· |
| C | 5852 hCoV-19/India/DL-MaxCov0031-CSIR-IGIB/2020\|EPI_ISL | ···················· | ···················· | ····················· | ···················· | ···················· | ···················· |
| C | 5853 hCoV-19/India/DL-MaxCov0032-CSIR-IGIB/2020\|EPI_ISL | ···················· | ···················· | ····················· | ···················· | ···················· | ···················· |
| C | 5854 hCoV-19/India/DL-MaxCov0033-CSIR-IGIB/2020\|EPI_ISL | ···················· | ···················· | ····················· | ···················· | ···················· | ···················· |
| C | 5855 hCoV-19/India/DL-MaxCov0034-CSIR-IGIB/2020\|EPI_ISL | ···················· | ···················· | ····················· | ···················· | ···················· | ···················· |
| C | 5856 hCoV-19/India/DL-MaxCov0035-CSIR-IGIB/2020\|EPI_ISL | ···················· | ···················· | ····················· | ···················· | ···················· | ···················· |
| C | 5857 hCoV-19/India/DL-MaxCov0036-CSIR-IGIB/2020\|EPI_ISL | ···················· | ···················· | ····················· | ···················· | ···················· | ···················· |
| C | 5858 hCoV-19/India/DL-MaxCov0037-CSIR-IGIB/2020\|EPI_ISL | ···················· | ···················· | ····················· | ···················· | ···················· | ···················· |
| C | 5859 hCoV-19/India/DL-MaxCov0038-CSIR-IGIB/2020\|EPI_ISL | ···················· | ···················· | ····················· | ···················· | ···················· | ···················· |
| C | 5860 hCoV-19/India/DL-MaxCov0039-CSIR-IGIB/2020\|EPI_ISL | ···················· | ···················· | ····················· | ···················· | ···················· | ···················· |
| C | 5861 hCoV-19/India/DL-MaxCov0040-CSIR-IGIB/2020\|EPI_ISL | ···················· | ···················· | ····················· | ···················· | ···················· | ···················· |
| C | 5862 hCoV-19/India/DL-MaxCov0041-CSIR-IGIB/2020\|EPI_ISL | ···················· | ···················· | ····················· | ···················· | ···················· | ···················· |
| C | 5863 hCoV-19/Netherlands/Gelderland_143/2020\|EPI_ISL_46 | ···················· | ···················· | ····················· | ···················· | ···················· | ···················· |
| C | 5864 hCoV-19/Netherlands/Gelderland_144/2020\|EPI_ISL_46 | ···················· | ···················· | ····················· | ···················· | ···················· | ···················· |
| C | 5865 hCoV-19/Netherlands/Gelderland_145/2020\|EPI_ISL_46 | ···················· | ···················· | ····················· | ···················· | ···················· | ···················· |
| C | 5866 hCoV-19/Netherlands/Gelderland_146/2020\|EPI_ISL_46 | ···················· | ···················· | ····················· | ···················· | ···················· | ···················· |
| C | 5867 hCoV-19/Netherlands/Gelderland_147/2020\|EPI_ISL_46 | ···················· | ···················· | ····················· | ···················· | ···················· | ···················· |
| C | 5868 hCoV-19/Netherlands/Gelderland_148/2020\|EPI_ISL_46 | ···················· | ···················· | ····················· | ···················· | ···················· | ···················· |
| C | 5869 hCoV-19/Netherlands/Gelderland_149/2020\|EPI_ISL_46 | ···················· | ···················· | ····················· | ···················· | ···················· | ···················· |
| C | 5870 hCoV-19/Netherlands/Gelderland_150/2020\|EPI_ISL_46 | ···················· | ···················· | ····················· | ···················· | ···················· | ···················· |
| C | 5871 hCoV-19/Netherlands/Gelderland_151/2020\|EPI_ISL_46 | ···················· | ···················· | ····················· | ···················· | ···················· | ···················· |
| C | 5872 hCoV-19/Netherlands/Gelderland_152/2020\|EPI_ISL_46 | ···················· | ···················· | ····················· | ···················· | ···················· | ···················· |
| C | 5873 hCoV-19/Netherlands/Gelderland_153/2020\|EPI_ISL_46 | ···················· | ···················· | ····················· | ···················· | ···················· | ···················· |
| C | 5874 hCoV-19/Netherlands/Gelderland_154/2020\|EPI_ISL_46 | ···················· | ···················· | ····················· | ···················· | ···················· | ···················· |
| C | 5875 hCoV-19/Netherlands/Gelderland_155/2020\|EPI_ISL_46 | ···················· | ···················· | ····················· | ···················· | ···················· | ···················· |
| C | 5876 hCoV-19/Netherlands/Gelderland_157/2020\|EPI_ISL_46 | ···················· | ···················· | ····················· | ···················· | ···················· | ···················· |
| C | 5877 hCoV-19/Netherlands/Gelderland_158/2020\|EPI_ISL_46 | ···················· | ···················· | ····················· | ···················· | ···················· | ···················· |
| C | 5878 hCoV-19/Netherlands/Gelderland_159/2020\|EPI_ISL_46 | ···················· | ···················· | ····················· | ···················· | ···················· | ···················· |
| C | 5879 hCoV-19/Netherlands/Gelderland_160/2020\|EPI_ISL_46 | ···················· | ···················· | ····················· | ···················· | ···················· | ···················· |
| C | 5880 hCoV-19/Netherlands/Gelderland_161/2020\|EPI_ISL_46 | ···················· | ···················· | ····················· | ···················· | ···················· | ···················· |
| C | 5881 hCoV-19/Netherlands/Gelderland_162/2020\|EPI_ISL_46 | ···················· | ···················· | ····················· | ···················· | ···················· | ···················· |
| C | 5882 hCoV-19/Netherlands/Gelderland_163/2020\|EPI_ISL_46 | ···················· | ···················· | ····················· | ···················· | ···················· | ···················· |
| C | 5883 hCoV-19/Netherlands/Gelderland_24/2020\|EPI_ISL_460 | ···················· | ···················· | ····················· | ···················· | ···················· | ···················· |
| C | 5884 hCoV-19/Netherlands/Gelderland_26/2020\|EPI_ISL_460 | ···················· | ···················· | ····················· | ···················· | ···················· | ···················· |
| C | 5885 hCoV-19/Netherlands/Gelderland_27/2020\|EPI_ISL_460 | ···················· | ···················· | ····················· | ···················· | ···················· | ···················· |
| C | 5886 hCoV-19/Netherlands/Gelderland_28/2020\|EPI_ISL_460 | ···················· | ···················· | ····················· | ···················· | ···················· | ···················· |
| C | 5887 hCoV-19/Netherlands/Gelderland_29/2020\|EPI_ISL_460 | ···················· | ···················· | ····················· | ···················· | ···················· | ···················· |
| C | 5888 hCoV-19/USA/NC-UNC-200414/2020\|EPI_ISL_524450\|2020 | ···················· | ···················· | ····················· | ···················· | ···················· | ···················· |
| C | 5889 hCoV-19/USA/PR-CDC-3578/2020\|EPI_ISL_447845\|2020-0 | ···················· | ···················· | ····················· | ···················· | ···················· | ···················· |
| C | 5890 hCoV-19/USA/VT-CDC-0303/2020\|EPI_ISL_447846\|2020-0 | ···················· | ···················· | ····················· | ···················· | ···················· | ···················· |
| C | 5891 hCoV-19/India/TG-CCMB-J045/2020\|EPI_ISL_447847\|202 | ···················· | ···················· | ····················· | ···················· | ···················· | ···················· |
| C | 5892 hCoV-19/India/TG-CCMB-J165/2020\|EPI_ISL_447848\|202 | ···················· | ···················· | ····················· | ···················· | ···················· | ···················· |
| C | 5893 hCoV-19/India/TG-CCMB-J107/2020\|EPI_ISL_447849\|202 | ···················· | ···················· | ····················· | ···················· | ···················· | ···················· |
| C | 5894 hCoV-19/India/TG-CCMB-J132/2020\|EPI_ISL_447850\|202 | ···················· | ···················· | ····················· | ···················· | ···················· | ···················· |
| C | 5895 hCoV-19/India/TG-CCMB-J223/2020\|EPI_ISL_447851\|202 | ···················· | ···················· | ····················· | ···················· | ···················· | ···················· |
| C | 5896 hCoV-19/India/TG-CCMB-J235/2020\|EPI_ISL_447852\|202 | ···················· | ···················· | ····················· | ···················· | ···················· | ···················· |
| C | 5897 hCoV-19/India/TG-CCMB-J247/2020\|EPI_ISL_447853\|202 | ···················· | ···················· | ····················· | ···················· | ···················· | ···················· |
| C | 5898 hCoV-19/India/TG-CCMB-J350/2020\|EPI_ISL_447854\|202 | ···················· | ···················· | ····················· | ···················· | ···················· | ···················· |
| C | 5899 hCoV-19/India/TG-CCMB-J318/2020\|EPI_ISL_447855\|202 | ···················· | ···················· | ····················· | ···················· | ···················· | ···················· |
| C | 5900 hCoV-19/India/TG-CCMB-J374/2020\|EPI_ISL_447856\|202 | ···················· | ···················· | ····················· | ···················· | ···················· | ···················· |
| C | 5901 hCoV-19/India/TG-CCMB-J390/2020\|EPI_ISL_447857\|202 | ···················· | ···················· | ····················· | ···················· | ···················· | ···················· |
| C | 5902 hCoV-19/India/TG-CCMB-J812/2020\|EPI_ISL_447858\|202 | ···················· | ···················· | ····················· | ···················· | ···················· | ···················· |
| C | 5903 hCoV-19/India/TG-CCMB-K771/2020\|EPI_ISL_447859\|202 | ···················· | ···················· | ····················· | ···················· | ···················· | ···················· |
| C | 5904 hCoV-19/India/TG-CCMB-K849/2020\|EPI_ISL_447860\|202 | ···················· | ···················· | ····················· | ···················· | ···················· | ···················· |
| C | 5905 hCoV-19/India/TG-CCMB-L149/2020\|EPI_ISL_447861\|202 | ···················· | ···················· | ····················· | ···················· | ···················· | ···················· |

| | | | | | | | | |
|---|---|---|---|---|---|---|---|---|
| C | 5906 | hCoV-19/India/TG-CCMB-J047/2020\|EPI_ISL_447862\|202 | .................... | .................... | .................... | .................... | .................... | .................... |
| C | 5907 | hCoV-19/India/TG-CCMB-K599/2020\|EPI_ISL_447863\|202 | .................... | .................... | .................... | .................... | .................... | .................... |
| C | 5908 | hCoV-19/India/TG-CCMB-L044/2020\|EPI_ISL_447864\|202 | .................... | .................... | .................... | .................... | .................... | .................... |
| C | 5909 | hCoV-19/India/TG-CCMB-K499/2020\|EPI_ISL_447865\|202 | .................... | .................... | .................... | .................... | .................... | .................... |
| C | 5910 | hCoV-19/India/TG-CCMB-K500/2020\|EPI_ISL_447866\|202 | .................... | .................... | .................... | .................... | .................... | .................... |
| C | 5911 | hCoV-19/USA/CA-CDC-0139/2020\|EPI_ISL_447886\|2020-0 | .................... | .................... | .................... | .................... | .................... | .................... |
| C | 5912 | hCoV-19/USA/CA-CZB-1136/2020\|EPI_ISL_447887\|2020-0 | .................... | .................... | .................... | .................... | .................... | .................... |
| C | 5913 | hCoV-19/USA/CA-CZB-1137/2020\|EPI_ISL_447888\|2020-0 | .................... | .................... | .................... | .................... | .................... | .................... |
| C | 5914 | hCoV-19/USA/CA-CZB-1138/2020\|EPI_ISL_447889\|2020-0 | .................... | .................... | .................... | .................... | .................... | .................... |
| C | 5915 | hCoV-19/USA/CA-CZB-1139/2020\|EPI_ISL_447890\|2020-0 | .................... | .................... | .................... | .................... | .................... | .................... |
| C | 5916 | hCoV-19/USA/CA-CZB-1140/2020\|EPI_ISL_447891\|2020-0 | .................... | .................... | .................... | .................... | .................... | .................... |
| C | 5917 | hCoV-19/USA/CA-CZB-1141/2020\|EPI_ISL_447892\|2020-0 | .................... | .................... | .................... | .................... | .................... | .................... |
| C | 5918 | hCoV-19/USA/CA-CZB-1142/2020\|EPI_ISL_447893\|2020-0 | .................... | .................... | .................... | .................... | .................... | .................... |
| C | 5919 | hCoV-19/USA/CA-CZB-1143/2020\|EPI_ISL_447894\|2020-0 | .................... | .................... | .................... | .................... | .................... | .................... |
| C | 5920 | hCoV-19/USA/CA-CZB-1144/2020\|EPI_ISL_447895\|2020-0 | .................... | .................... | .................... | .................... | .................... | .................... |
| C | 5921 | hCoV-19/USA/CA-CZB-1145/2020\|EPI_ISL_447896\|2020-0 | .................... | .................... | .................... | .................... | .................... | .................... |
| C | 5922 | hCoV-19/Bangladesh/CHRF-01/2020\|EPI_ISL_447899\|202 | .................... | .................... | .................... | .................... | .................... | .................... |
| C | 5923 | hCoV-19/Australia/SAP063/2020\|EPI_ISL_451102\|2020- | .................... | .................... | .................... | .................... | .................... | .................... |
| C | 5924 | hCoV-19/Australia/SAP065/2020\|EPI_ISL_451103\|2020- | .................... | .................... | .................... | .................... | .................... | .................... |
| C | 5925 | hCoV-19/Australia/SAP066/2020\|EPI_ISL_451104\|2020- | .................... | .................... | .................... | .................... | .................... | .................... |
| C | 5926 | hCoV-19/Australia/SAP067/2020\|EPI_ISL_451105\|2020- | .................... | .................... | .................... | .................... | .................... | .................... |
| C | 5927 | hCoV-19/Australia/SAP068/2020\|EPI_ISL_451106\|2020- | .................... | .................... | .................... | .................... | .................... | .................... |
| C | 5928 | hCoV-19/Australia/SAP069/2020\|EPI_ISL_451107\|2020- | .................... | .................... | .................... | .................... | .................... | .................... |
| C | 5929 | hCoV-19/Australia/SAP070/2020\|EPI_ISL_451108\|2020- | .................... | .................... | .................... | .................... | .................... | .................... |
| C | 5930 | hCoV-19/Australia/SAP073/2020\|EPI_ISL_451110\|2020- | .................... | .................... | .................... | .................... | .................... | .................... |
| C | 5931 | hCoV-19/Australia/SAP074/2020\|EPI_ISL_451111\|2020- | .................... | .................... | .................... | .................... | .................... | .................... |
| C | 5935 | hCoV-19/Spain/AN-IBV-001935/2020\|EPI_ISL_452441\|20 | .................... | .................... | .................... | .................... | .................... | .................... |
| C | 5936 | hCoV-19/England/SHEF-C2AC0/2020\|EPI_ISL_453701\|202 | .................... | .................... | .................... | .................... | .................... | .................... |
| C | 5937 | hCoV-19/England/SHEF-C2BDC/2020\|EPI_ISL_453703\|202 | .................... | .................... | .................... | .................... | .................... | .................... |
| C | 5938 | hCoV-19/England/SHEF-C2D6D/2020\|EPI_ISL_453704\|202 | .................... | .................... | .................... | .................... | .................... | .................... |
| C | 5939 | hCoV-19/England/SHEF-C33C4/2020\|EPI_ISL_453705\|202 | .................... | .................... | .................... | .................... | .................... | .................... |
| C | 5940 | hCoV-19/England/SHEF-C3C23/2020\|EPI_ISL_453706\|202 | .................... | .................... | .................... | .................... | .................... | .................... |
| C | 5941 | hCoV-19/England/SHEF-C3C7E/2020\|EPI_ISL_453707\|202 | .................... | .................... | .................... | .................... | .................... | .................... |
| C | 5942 | hCoV-19/England/SHEF-C3FA2/2020\|EPI_ISL_453708\|202 | .................... | .................... | .................... | .................... | .................... | .................... |
| C | 5943 | hCoV-19/England/SHEF-C43D2/2020\|EPI_ISL_453709\|202 | .................... | .............A...... | .................... | .................... | .................... | .................... |
| C | 5944 | hCoV-19/England/SHEF-C4615/2020\|EPI_ISL_453710\|202 | .................... | .................... | .................... | .................... | .................... | .................... |
| C | 5945 | hCoV-19/England/SHEF-C47B8/2020\|EPI_ISL_453711\|202 | .................... | .................... | .................... | .................... | .................... | .................... |
| C | 5946 | hCoV-19/England/SHEF-C4C7D/2020\|EPI_ISL_453712\|202 | .................... | .................... | .................... | .................... | .................... | .................... |
| C | 5947 | hCoV-19/England/SHEF-C4D6B/2020\|EPI_ISL_453713\|202 | .................... | .................... | .................... | .................... | .................... | .................... |
| C | 5948 | hCoV-19/England/SHEF-C51B9/2020\|EPI_ISL_453714\|202 | .................... | .................... | .................... | .................... | .................... | .................... |
| C | 5949 | hCoV-19/England/SHEF-C5614/2020\|EPI_ISL_453715\|202 | .................... | .................... | .................... | .................... | .................... | .................... |
| C | 5950 | hCoV-19/England/SHEF-C5C4F/2020\|EPI_ISL_453717\|202 | .................... | .................... | .................... | .................... | .................... | .................... |
| C | 5951 | hCoV-19/England/SHEF-C65DA/2020\|EPI_ISL_453719\|202 | .................... | .................... | .................... | .................... | .................... | .................... |
| C | 5952 | hCoV-19/England/SHEF-D1369/2020\|EPI_ISL_453765\|202 | .................... | .................... | .................... | .................... | .................... | .................... |
| C | 5953 | hCoV-19/England/SHEF-D18B5/2020\|EPI_ISL_453766\|202 | .................... | .................... | .................... | .................... | .................... | .................... |
| C | 5954 | hCoV-19/England/SHEF-D19B2/2020\|EPI_ISL_453767\|202 | .................... | .................... | .................... | .................... | .................... | .................... |
| C | 5955 | hCoV-19/England/SHEF-D1D3E/2020\|EPI_ISL_453768\|202 | .................... | .................... | .................... | .................... | .................... | .................... |
| C | 5956 | hCoV-19/England/SHEF-D1F65/2020\|EPI_ISL_453769\|202 | .................... | .................... | .................... | .................... | .................... | .................... |
| C | 5957 | hCoV-19/England/SHEF-D2104/2020\|EPI_ISL_453770\|202 | .................... | .................... | .................... | .................... | .................... | .................... |
| C | 5958 | hCoV-19/England/SHEF-D2438/2020\|EPI_ISL_453771\|202 | .................... | .................... | .................... | .................... | .................... | .................... |
| C | 5959 | hCoV-19/England/SHEF-D2A36/2020\|EPI_ISL_453772\|202 | .................... | .................... | .................... | .................... | .................... | .................... |
| C | 5960 | hCoV-19/England/SHEF-D2AFA/2020\|EPI_ISL_453773\|202 | .................... | .................... | .................... | .................... | .................... | .................... |
| C | 5961 | hCoV-19/England/EXET-13611C/2020\|EPI_ISL_457200\|20 | .................... | .................... | .................... | .................... | .................... | .................... |
| C | 5962 | hCoV-19/England/EXET-13613A/2020\|EPI_ISL_457201\|20 | .................... | .................... | .................... | .................... | .................... | .................... |
| C | 5963 | hCoV-19/England/EXET-136167/2020\|EPI_ISL_457204\|20 | .................... | .................... | .................... | .................... | .................... | .................... |
| C | 5965 | hCoV-19/England/EXET-1362EC/2020\|EPI_ISL_457217\|20 | .................... | .................... | .................... | .................... | .................... | .................... |
| C | 5966 | hCoV-19/England/EXET-136307/2020\|EPI_ISL_457218\|20 | .................... | .................... | .................... | .................... | .................... | .................... |
| C | 5968 | hCoV-19/England/EXET-136334/2020\|EPI_ISL_457221\|20 | .................... | .................... | .................... | .................... | .................... | .................... |
| C | 5969 | hCoV-19/England/EXET-136352/2020\|EPI_ISL_457222\|20 | .................... | .................... | .................... | .................... | .................... | .................... |
| C | 5971 | hCoV-19/England/EXET-1363CB/2020\|EPI_ISL_457224\|20 | .................... | .................... | .................... | .................... | .................... | .................... |
| C | 5972 | hCoV-19/England/EXET-1363DA/2020\|EPI_ISL_457225\|20 | .................... | .................... | .................... | .................... | .................... | .................... |
| C | 5979 | hCoV-19/England/EXET-1368EA/2020\|EPI_ISL_457239\|20 | .................... | .................... | .................... | .................... | .................... | .................... |
| C | 5980 | hCoV-19/England/EXET-1368F9/2020\|EPI_ISL_457240\|20 | .................... | .................... | .................... | .................... | .................... | .................... |
| C | 5981 | hCoV-19/England/EXET-136914/2020\|EPI_ISL_457241\|20 | .................... | .................... | .................... | .................... | .................... | .................... |
| C | 5983 | hCoV-19/England/EXET-136A7B/2020\|EPI_ISL_457245\|20 | .................... | .................... | .................... | .................... | .N.................. | .................... |
| C | 5984 | hCoV-19/England/EXET-136A99/2020\|EPI_ISL_457246\|20 | .................... | .................... | .................... | .................... | .................... | .................... |
| C | 5985 | hCoV-19/England/EXET-136AC6/2020\|EPI_ISL_457247\|20 | .................... | .................... | .................... | .................... | .................... | .................... |
| C | 5987 | hCoV-19/England/EXET-136B4B/2020\|EPI_ISL_457249\|20 | .................... | .................... | .................... | .................... | .................... | .................... |
| C | 5988 | hCoV-19/England/EXET-136B69/2020\|EPI_ISL_457250\|20 | .................... | .................... | .................... | .................... | .................... | .................... |
| C | 5989 | hCoV-19/England/EXET-136B78/2020\|EPI_ISL_457251\|20 | .................... | .................... | .................... | .................... | .................... | .................... |
| C | 5990 | hCoV-19/England/EXET-136B87/2020\|EPI_ISL_457252\|20 | .................... | .................... | .................... | .................... | .................... | .................... |
| C | 5991 | hCoV-19/England/EXET-136B96/2020\|EPI_ISL_457253\|20 | .................... | .................... | .................... | .................... | .................... | .................... |
| C | 5992 | hCoV-19/England/EXET-136BC3/2020\|EPI_ISL_457254\|20 | .................... | .................... | .................... | .................... | .................... | .................... |
| C | 5993 | hCoV-19/England/EXET-136BF0/2020\|EPI_ISL_457255\|20 | .................... | ...N................ | .................... | .................... | .................... | .................... |
| C | 5994 | hCoV-19/England/EXET-136C0C/2020\|EPI_ISL_457256\|20 | .................... | .................... | .................... | .................... | .................... | .................... |
| C | 5995 | hCoV-19/England/EXET-136C57/2020\|EPI_ISL_457258\|20 | .................... | .................... | .................... | .................... | .................... | .................... |
| C | 5996 | hCoV-19/England/LOND-D5884/2020\|EPI_ISL_457270\|202 | .................... | .................... | .................... | .................... | .................... | .................... |
| C | 5997 | hCoV-19/England/LOND-D5893/2020\|EPI_ISL_457271\|202 | .................... | ..................C. | .................... | .................... | .................... | .................... |

| | | | | | | | | |
|---|---|---|---|---|---|---|---|---|
| C | 5998 hCoV-19/England/LOND-D58B1/2020\|EPI_ISL_457272\|202 | ···················· | ···················· | ···················· | ···················· | ···················· | ···················· | |
| C | 5999 hCoV-19/England/LOND-D58C0/2020\|EPI_ISL_457273\|202 | ···················· | ···················· | ···················· | ···················· | ···················· | ···················· | |
| C | 6000 hCoV-19/England/LOND-D58DF/2020\|EPI_ISL_457274\|202 | ···················· | ···················· | ···················· | ···················· | ···················· | ···················· | |
| C | 6001 hCoV-19/England/LOND-D58EE/2020\|EPI_ISL_457275\|202 | ···················· | ···················· | ···················· | ···················· | ···················· | ···················· | |
| C | 6002 hCoV-19/England/LOND-D5909/2020\|EPI_ISL_457277\|202 | ···················· | ···················· | ···················· | ···················· | ···T················ | ···················· | |
| C | 6003 hCoV-19/England/LOND-D5918/2020\|EPI_ISL_457278\|202 | ···················· | ···················· | ···················· | ···················· | ···················· | ···················· | |
| C | 6004 hCoV-19/England/LOND-D5936/2020\|EPI_ISL_457280\|202 | ···················· | ···················· | ···················· | ···················· | ···················· | ···················· | |
| C | 6005 hCoV-19/England/LOND-D5945/2020\|EPI_ISL_457281\|202 | ···················· | ···················· | ···················· | ···················· | ···················· | ···················· | |
| C | 6006 hCoV-19/England/LOND-D5981/2020\|EPI_ISL_457283\|202 | ···················· | ···················· | ···················· | ···················· | ···················· | ···················· | |
| C | 6007 hCoV-19/England/LOND-D5990/2020\|EPI_ISL_457284\|202 | ···················· | ···················· | ···················· | ···················· | ···················· | ···················· | |
| C | 6008 hCoV-19/England/LOND-D59AF/2020\|EPI_ISL_457285\|202 | ···················· | ···················· | ···················· | ···················· | ···················· | ···················· | |
| C | 6009 hCoV-19/England/LOND-D59BE/2020\|EPI_ISL_457286\|202 | ···················· | ···················· | ···················· | ···················· | ···················· | ···················· | |
| C | 6010 hCoV-19/England/LOND-D59CD/2020\|EPI_ISL_457287\|202 | ···················· | ···················· | ···················· | ···················· | ···················· | ···················· | |
| C | 6011 hCoV-19/England/LOND-D59EB/2020\|EPI_ISL_457288\|202 | ···················· | ··················C· | ···················· | ···················· | ···················· | ···················· | 1 |
| C | 6012 hCoV-19/England/LOND-D59FA/2020\|EPI_ISL_457289\|202 | ···················· | ···················· | ···················· | ···················· | ···················· | ···················· | |
| C | 6013 hCoV-19/England/LOND-D5A06/2020\|EPI_ISL_457290\|202 | ···················· | ···················· | ···················· | ···················· | ···················· | ···················· | |
| C | 6014 hCoV-19/England/LOND-D5A15/2020\|EPI_ISL_457291\|202 | ···················· | ···················· | ···················· | ···················· | ···················· | ···················· | |
| C | 6015 hCoV-19/England/LOND-D5A33/2020\|EPI_ISL_457292\|202 | ···················· | ···················· | ···················· | ···················· | ···················· | ···················· | |
| C | 6016 hCoV-19/England/LOND-D5A42/2020\|EPI_ISL_457293\|202 | ···················· | ···················· | ···················· | ···················· | ···················· | ···················· | |
| C | 6017 hCoV-19/England/LOND-D5A60/2020\|EPI_ISL_457294\|202 | ···················· | ···················· | ···················· | ···················· | ···················· | ···················· | |
| C | 6018 hCoV-19/England/LOND-D5A7F/2020\|EPI_ISL_457295\|202 | ···················· | ···················· | ···················· | ···················· | ···················· | ···················· | |
| C | 6019 hCoV-19/England/LOND-D5A9D/2020\|EPI_ISL_457296\|202 | ···················· | ···················· | ···················· | ···················· | ···················· | ···················· | |
| C | 6020 hCoV-19/England/LOND-D5ACA/2020\|EPI_ISL_457297\|202 | ···················· | ···················· | ···················· | ···················· | ···················· | ···················· | |
| C | 6021 hCoV-19/England/LOND-D5AE8/2020\|EPI_ISL_457298\|202 | ···················· | ···················· | ···················· | ···················· | ···················· | ···················· | |
| C | 6022 hCoV-19/England/LOND-D5AF7/2020\|EPI_ISL_457299\|202 | ···················· | ···················· | ···················· | ···················· | ···················· | ···················· | |
| C | 6023 hCoV-19/England/BRIS-129D72/2020\|EPI_ISL_458500\|20 | ···················· | ···················· | ···················· | ···················· | ···················· | ···················· | |
| C | 6024 hCoV-19/England/BRIS-129D36/2020\|EPI_ISL_458501\|20 | ···················· | ···················· | ···················· | ···················· | ···················· | ···················· | |
| C | 6025 hCoV-19/England/BRIS-12ACCF/2020\|EPI_ISL_458502\|20 | ···················· | ···················· | ···················· | ···················· | ···················· | ···················· | |
| C | 6026 hCoV-19/England/BRIS-1298BD/2020\|EPI_ISL_458503\|20 | ···················· | ···················· | ···················· | ···················· | ···················· | ···················· | |
| C | 6027 hCoV-19/England/BRIS-129E51/2020\|EPI_ISL_458507\|20 | ···················· | ···················· | ···················· | ···················· | ···················· | ···················· | |
| C | 6028 hCoV-19/England/BRIS-12A1EE/2020\|EPI_ISL_458509\|20 | ···················· | ···················· | ···················· | ···················· | ···················· | ···················· | |
| C | 6029 hCoV-19/England/BRIS-12AC29/2020\|EPI_ISL_458510\|20 | ···················· | ···················· | ···················· | ···················· | ···················· | ···················· | |
| C | 6030 hCoV-19/England/BRIS-12A2EB/2020\|EPI_ISL_458511\|20 | ···················· | ···················· | ···················· | ···················· | ···················· | ···················· | |
| C | 6031 hCoV-19/England/BRIS-12A0E1/2020\|EPI_ISL_458512\|20 | ···················· | ···················· | ···················· | ···················· | ···················· | ···················· | |
| C | 6032 hCoV-19/England/BRIS-12AF6C/2020\|EPI_ISL_458513\|20 | ···················· | ···················· | ···················· | ···················· | ···················· | ···················· | |
| C | 6033 hCoV-19/England/BRIS-12AAA7/2020\|EPI_ISL_458515\|20 | ···················· | ···················· | ···················· | ···················· | ···················· | ···················· | |
| C | 6034 hCoV-19/England/CAMB-1B1595/2020\|EPI_ISL_458529\|20 | ···················· | ···················· | ···················· | ···················· | ···················· | ···················· | |
| C | 6035 hCoV-19/England/CAMB-1B143E/2020\|EPI_ISL_458533\|20 | ···················· | ···················· | ···················· | ···················· | ···················· | ···················· | |
| C | 6036 hCoV-19/England/CAMB-1B1DCA/2020\|EPI_ISL_458552\|20 | ···················· | ···················· | ···················· | ···················· | ···················· | ···················· | |
| C | 6037 hCoV-19/England/CAMB-1B1498/2020\|EPI_ISL_458556\|20 | ···················· | ···················· | ···················· | ···················· | ···················· | ···················· | |
| C | 6038 hCoV-19/England/CAMB-1B15E0/2020\|EPI_ISL_458559\|20 | ···················· | ···················· | ···················· | ···················· | ···················· | ···················· | |
| C | 6039 hCoV-19/England/CAMB-1B13C8/2020\|EPI_ISL_458576\|20 | ···················· | ···················· | ···················· | ···················· | ···················· | ···················· | |
| C | 6040 hCoV-19/England/NORT-288F10/2020\|EPI_ISL_458579\|20 | ···················· | ···················· | ···················· | ···················· | ···················· | ···················· | |
| C | 6041 hCoV-19/England/NORT-288F3E/2020\|EPI_ISL_458580\|20 | ···················· | ···················· | ···················· | ···················· | ···················· | ···················· | |
| C | 6042 hCoV-19/England/NORT-289708/2020\|EPI_ISL_458581\|20 | ···················· | ···················· | ···················· | ···················· | ···················· | ···················· | |
| C | 6043 hCoV-19/England/NORT-289207/2020\|EPI_ISL_458583\|20 | ···················· | ···················· | ···················· | ···················· | ···················· | ···················· | |
| C | 6044 hCoV-19/England/NORT-2891CE/2020\|EPI_ISL_458584\|20 | ···················· | ···················· | ···················· | ···················· | ···················· | ···················· | |
| C | 6045 hCoV-19/England/NORT-2892E9/2020\|EPI_ISL_458588\|20 | ···················· | ···················· | ···················· | ···················· | ···················· | ···················· | |
| C | 6046 hCoV-19/England/NORT-289841/2020\|EPI_ISL_458589\|20 | ···················· | ···················· | ···················· | ···················· | ···················· | ···················· | |
| C | 6047 hCoV-19/England/NORT-288DBC/2020\|EPI_ISL_458591\|20 | ···················· | ···················· | ···················· | ···················· | ···················· | ···················· | |
| C | 6048 hCoV-19/England/NORT-2895D1/2020\|EPI_ISL_458593\|20 | ···················· | ···················· | ···················· | ···················· | ···················· | ···················· | |
| C | 6049 hCoV-19/England/NORT-28953B/2020\|EPI_ISL_458597\|20 | ···················· | ···················A | ···················· | ···················· | ···················· | ···················· | 1 |
| C | 6050 hCoV-19/Chile/RM-CMM-26347/2020\|EPI_ISL_459856\|202 | ···················· | ···················· | ···················· | ···················· | ···················· | ···················· | |
| C | 6051 hCoV-19/Chile/RM-CMM-27991/2020\|EPI_ISL_459857\|202 | ···················· | ···················· | ···················· | ···················· | ···················· | ···················· | |
| C | 6052 hCoV-19/Chile/RM-CMM-26639/2020\|EPI_ISL_459858\|202 | ···················· | ···················· | ···················· | ···················· | ···················· | ···················· | |
| C | 6053 hCoV-19/Chile/RM-CMM-32205/2020\|EPI_ISL_459859\|202 | ···················· | ···················· | ···················· | ···················· | ···················· | ···················· | |
| C | 6054 hCoV-19/Chile/RM-CMM-29192/2020\|EPI_ISL_459860\|202 | ···················· | ···················· | ···················· | ···················· | ···················· | ···················· | |
| C | 6055 hCoV-19/Chile/MA-CMM-31600/2020\|EPI_ISL_459861\|202 | ···················· | ···················· | ···················· | ···················· | ···················· | ···················· | |
| C | 6056 hCoV-19/Chile/RM-CMM-25669/2020\|EPI_ISL_459862\|202 | ···················· | ···················· | ···················· | ···················· | ···················· | ···················· | |
| C | 6057 hCoV-19/Chile/ML-CMM-25343/2020\|EPI_ISL_459863\|202 | ···················· | ···················· | ···················· | ···················· | ···················· | ···················· | |
| C | 6058 hCoV-19/Canada/ON_QGLO-01/2020\|EPI_ISL_459866\|2020 | ···················· | ···················· | ···················· | ···················· | ···················· | ···················· | |
| C | 6059 hCoV-19/Canada/ON_QGLO-02/2020\|EPI_ISL_459867\|2020 | ···················· | ···················· | ···················· | ···················· | ···················· | ···················· | |
| C | 6060 hCoV-19/Canada/ON_QGLO-023/2020\|EPI_ISL_459879\|202 | ···················· | ···················· | ···················· | ···················· | ···················· | ···················· | |
| C | 6061 hCoV-19/Canada/ON_QGLO-024/2020\|EPI_ISL_459880\|202 | ···················· | ···················· | ···················· | ···················· | ···················· | ···················· | |
| C | 6062 hCoV-19/Canada/ON_QGLO-025/2020\|EPI_ISL_459881\|202 | ···················· | ···················· | ···················· | ···················· | ···················· | ···················· | |
| C | 6063 hCoV-19/Canada/ON_QGLO-026/2020\|EPI_ISL_459882\|202 | ···················· | ···················· | ···················· | ···················· | ···················· | ···················· | |
| C | 6064 hCoV-19/Canada/ON_QGLO-029/2020\|EPI_ISL_459883\|202 | ···················· | ···················· | ···················· | ···················· | ···················· | ···················· | |
| C | 6065 hCoV-19/Canada/ON_QGLO-034/2020\|EPI_ISL_459885\|202 | ···················· | ···················· | ···················· | ···················· | ···················· | ···················· | |
| C | 6066 hCoV-19/Canada/ON_QGLO-035/2020\|EPI_ISL_459886\|202 | ···················· | ···················· | ···················· | ···················· | ···················· | ···················· | |
| C | 6067 hCoV-19/Canada/ON_QGLO-036/2020\|EPI_ISL_459887\|202 | ···················· | ···················· | ···················· | ···················· | ···················· | ···················· | |
| C | 6068 hCoV-19/Canada/ON_QGLO-037/2020\|EPI_ISL_459888\|202 | ···················· | ···················· | ···················· | ···················· | ···················· | ···················· | |
| C | 6069 hCoV-19/Canada/ON_QGLO-038/2020\|EPI_ISL_459889\|202 | ···················· | ···················· | ···················· | ···················· | ···················· | ···················· | |
| C | 6070 hCoV-19/Canada/ON_QGLO-039/2020\|EPI_ISL_459890\|202 | ···················· | ···················· | ···················· | ···················· | ···················· | ···················· | |
| C | 6071 hCoV-19/Canada/ON_QGLO-041/2020\|EPI_ISL_459891\|202 | ···················· | ···················· | ···················· | ···················· | ···················· | ···················· | |
| C | 6072 hCoV-19/Canada/ON_QGLO-042/2020\|EPI_ISL_459892\|202 | ···················· | ···················· | ···················· | ···················· | ···················· | ···················· | |
| C | 6073 hCoV-19/Luxembourg/LNS0681882/2020\|EPI_ISL_459893\| | ···················· | ···················· | ···················· | ···················· | ···················· | ···················· | |
| C | 6074 hCoV-19/Luxembourg/LNS1646752/2020\|EPI_ISL_459894\| | ···················· | ···················· | ···················· | ···················· | ···················· | ···················· | |
| C | 6075 hCoV-19/Luxembourg/LNS3660996/2020\|EPI_ISL_459896\| | ···················· | ···················· | ···················· | ···················· | ···················· | ···················· | |

| | | | | | | | | |
|---|---|---|---|---|---|---|---|---|
| C | 6076 | hCoV-19/Luxembourg/LNS3923316/2020\|EPI_ISL_459898\| | ···················· | ···················· | ····················· | ···················· | ···················· | ···················· |
| C | 6077 | hCoV-19/Luxembourg/LNS5507695/2020\|EPI_ISL_459899\| | ···················· | ···················· | ····················· | ···················· | ···················· | ···················· |
| C | 6078 | hCoV-19/Netherlands/Overijssel_21/2020\|EPI_ISL_460 | ···················· | ···················· | ····················· | ···················· | ···················· | ···················· |
| C | 6079 | hCoV-19/Netherlands/Overijssel_22/2020\|EPI_ISL_460 | ···················· | ···················· | ····················· | ···················· | ···················· | ···················· |
| C | 6080 | hCoV-19/Netherlands/Overijssel_24/2020\|EPI_ISL_460 | ···················· | ···················· | ····················· | ···················· | ···················· | ···················· |
| C | 6081 | hCoV-19/Netherlands/Overijssel_25/2020\|EPI_ISL_460 | ···················· | ···················· | ····················· | ···················· | ···················· | ···················· |
| C | 6082 | hCoV-19/Netherlands/Overijssel_26/2020\|EPI_ISL_460 | ···················· | ···················· | ····················· | ···················· | ···················· | ···················· |
| C | 6083 | hCoV-19/Netherlands/Overijssel_27/2020\|EPI_ISL_460 | ···················· | ···················· | ····················· | ···················· | ···················· | ···················· |
| C | 6084 | hCoV-19/Netherlands/Overijssel_28/2020\|EPI_ISL_460 | ···················· | ···················· | ····················· | ···················· | ···················· | ···················· |
| C | 6085 | hCoV-19/Netherlands/Overijssel_29/2020\|EPI_ISL_460 | ···················· | ···················· | ····················· | ···················· | ···················· | ···················· |
| C | 6086 | hCoV-19/Netherlands/Overijssel_3/2020\|EPI_ISL_4608 | ···················· | ···················· | ····················· | ···················· | ···················· | ···················· |
| C | 6087 | hCoV-19/Netherlands/Overijssel_30/2020\|EPI_ISL_460 | ···················· | ···················· | ····················· | ···················· | ···················· | ···················· |
| C | 6088 | hCoV-19/Netherlands/Overijssel_31/2020\|EPI_ISL_460 | ···················· | ···················· | ····················· | ···················· | ···················· | ···················· |
| C | 6089 | hCoV-19/Netherlands/Overijssel_32/2020\|EPI_ISL_460 | ···················· | ···················· | ····················· | ···················· | ···················· | ···················· |
| C | 6090 | hCoV-19/Netherlands/Overijssel_4/2020\|EPI_ISL_4608 | ···················· | ···················· | ····················· | ···················· | ···················· | ···················· |
| C | 6091 | hCoV-19/Netherlands/Overijssel_5/2020\|EPI_ISL_4608 | ···················· | ···················· | ····················· | ···················· | ···················· | ···················· |
| C | 6092 | hCoV-19/Netherlands/Overijssel_6/2020\|EPI_ISL_4608 | ···················· | ···················· | ····················· | ···················· | ···················· | ···················· |
| C | 6093 | hCoV-19/Netherlands/Overijssel_7/2020\|EPI_ISL_4608 | ···················· | ···················· | ····················· | ···················· | ···················· | ···················· |
| C | 6094 | hCoV-19/Netherlands/Overijssel_8/2020\|EPI_ISL_4608 | ···················· | ···················· | ····················· | ···················· | ···················· | ···················· |
| C | 6095 | hCoV-19/Netherlands/Overijssel_9/2020\|EPI_ISL_4608 | ···················· | ···················· | ····················· | ···················· | ···················· | ···················· |
| C | 6096 | hCoV-19/Netherlands/Utrecht_26/2020\|EPI_ISL_460829 | ···················· | ··········A········· | ····················· | ···················· | ···················· | ···················· |
| C | 6097 | hCoV-19/Netherlands/Utrecht_27/2020\|EPI_ISL_460830 | ···················· | ···················· | ····················· | ···················· | ···················· | ···················· |
| C | 6098 | hCoV-19/Netherlands/Utrecht_28/2020\|EPI_ISL_460831 | ···················· | ···················· | ····················· | ···················· | ···················· | ···················· |
| C | 6099 | hCoV-19/Netherlands/Utrecht_29/2020\|EPI_ISL_460832 | ···················· | ···················· | ····················· | ···················· | ···················· | ···················· |
| C | 6100 | hCoV-19/Netherlands/Flevoland_10/2020\|EPI_ISL_4608 | ···················· | ···················· | ····················· | ···················· | ···················· | ···················· |
| C | 6101 | hCoV-19/Netherlands/Flevoland_11/2020\|EPI_ISL_4608 | ···················· | ···················· | ····················· | ···················· | ···················· | ···················· |
| C | 6102 | hCoV-19/Netherlands/Flevoland_12/2020\|EPI_ISL_4608 | ···················· | ···················· | ····················· | ···················· | ···················· | ···················· |
| C | 6103 | hCoV-19/Netherlands/Flevoland_13/2020\|EPI_ISL_4608 | ···················· | ···················· | ····················· | ···················· | ···················· | ···················· |
| C | 6104 | hCoV-19/Netherlands/Flevoland_14/2020\|EPI_ISL_4608 | ···················· | ···················· | ····················· | ···················· | ···················· | ···················· |
| C | 6105 | hCoV-19/Netherlands/Flevoland_15/2020\|EPI_ISL_4608 | ···················· | ···················· | ····················· | ···················· | ···················· | ···················· |
| C | 6106 | hCoV-19/Netherlands/Flevoland_16/2020\|EPI_ISL_4608 | ···················· | ···················· | ····················· | ···················· | ···················· | ···················· |
| C | 6107 | hCoV-19/Netherlands/Flevoland_17/2020\|EPI_ISL_4608 | ···················· | ···················· | ····················· | ···················· | ···················· | ···················· |
| C | 6108 | hCoV-19/Netherlands/Flevoland_18/2020\|EPI_ISL_4608 | ···················· | ···················· | ····················· | ···················· | ···················· | ···················· |
| C | 6109 | hCoV-19/Netherlands/Flevoland_8/2020\|EPI_ISL_46084 | ···················· | ···················· | ····················· | ···················· | ···················· | ···················· |
| C | 6110 | hCoV-19/Netherlands/Flevoland_9/2020\|EPI_ISL_46084 | ···················· | ···················· | ····················· | ···················· | ···················· | ···················· |
| C | 6111 | hCoV-19/Netherlands/Friesland_29/2020\|EPI_ISL_4608 | ···················· | ···················· | ····················· | ···················· | ···················· | ···················· |
| C | 6112 | hCoV-19/Netherlands/Friesland_30/2020\|EPI_ISL_4608 | ···················· | ···················· | ····················· | ···················· | ···················· | ···················· |
| C | 6113 | hCoV-19/Netherlands/Friesland_32/2020\|EPI_ISL_4608 | ···················· | ···················· | ····················· | ···················· | ···················· | ···················· |
| C | 6114 | hCoV-19/Netherlands/Friesland_34/2020\|EPI_ISL_4608 | ···················· | ···················· | ····················· | ···················· | ···················· | ···················· |
| C | 6115 | hCoV-19/Netherlands/Friesland_45/2020\|EPI_ISL_4608 | ···················· | ···················· | ····················· | ···················· | ···················· | ···················· |
| C | 6116 | hCoV-19/Netherlands/Gelderland_100/2020\|EPI_ISL_46 | ···················· | ···················· | ····················· | ···················· | ···················· | ···················· |
| C | 6117 | hCoV-19/Netherlands/Gelderland_101/2020\|EPI_ISL_46 | ···················· | ···················· | ····················· | ···················· | ···················· | ···················· |
| C | 6118 | hCoV-19/Netherlands/Gelderland_102/2020\|EPI_ISL_46 | ···················· | ···················· | ····················· | ···················· | ···················· | ···················· |
| C | 6119 | hCoV-19/Netherlands/Gelderland_103/2020\|EPI_ISL_46 | ···················· | ···················· | ····················· | ···················· | ···················· | ···················· |
| C | 6120 | hCoV-19/Netherlands/Gelderland_104/2020\|EPI_ISL_46 | ···················· | ···················· | ····················· | ···················· | ···················· | ···················· |
| C | 6121 | hCoV-19/Netherlands/Gelderland_107/2020\|EPI_ISL_46 | ···················· | ···················· | ····················· | ···················· | ···················· | ···················· |
| C | 6122 | hCoV-19/Netherlands/Gelderland_108/2020\|EPI_ISL_46 | ···················· | ···················· | ····················· | ···················· | ···················· | ···················· |
| C | 6123 | hCoV-19/Netherlands/Gelderland_109/2020\|EPI_ISL_46 | ···················· | ···················· | ····················· | ···················· | ···················· | ···················· |
| C | 6124 | hCoV-19/Netherlands/Gelderland_110/2020\|EPI_ISL_46 | ···················· | ···················· | ····················· | ···················· | ···················· | ···················· |
| C | 6125 | hCoV-19/Netherlands/Gelderland_111/2020\|EPI_ISL_46 | ···················· | ···················· | ····················· | ···················· | ···················· | ···················· |
| C | 6126 | hCoV-19/Netherlands/Gelderland_112/2020\|EPI_ISL_46 | ···················· | ···················· | ····················· | ···················· | ···················· | ···················· |
| C | 6127 | hCoV-19/Netherlands/Gelderland_113/2020\|EPI_ISL_46 | ···················· | ···················· | ····················· | ···················· | ···················· | ···················· |
| C | 6128 | hCoV-19/Netherlands/Gelderland_114/2020\|EPI_ISL_46 | ···················· | ···················· | ····················· | ···················· | ···················· | ···················· |
| C | 6129 | hCoV-19/Netherlands/Gelderland_115/2020\|EPI_ISL_46 | ···················· | ···················· | ····················· | ···················· | ···················· | ···················· |
| C | 6130 | hCoV-19/Netherlands/Gelderland_116/2020\|EPI_ISL_46 | ···················· | ···················· | ····················· | ···················· | ···················· | ···················· |
| C | 6131 | hCoV-19/Netherlands/Gelderland_117/2020\|EPI_ISL_46 | ···················· | ···················· | ····················· | ···················· | ···················· | ···················· |
| C | 6132 | hCoV-19/Netherlands/Gelderland_118/2020\|EPI_ISL_46 | ···················· | ···················· | ····················· | ···················· | ···················· | ···················· |
| C | 6133 | hCoV-19/Netherlands/Gelderland_119/2020\|EPI_ISL_46 | ···················· | ···················· | ····················· | ···················· | ···················· | ···················· |
| C | 6134 | hCoV-19/Netherlands/Gelderland_120/2020\|EPI_ISL_46 | ···················· | ···················· | ····················· | ···················· | ···················· | ···················· |
| C | 6135 | hCoV-19/Netherlands/Gelderland_121/2020\|EPI_ISL_46 | ···················· | ···················· | ····················· | ···················· | ···················· | ···················· |
| C | 6136 | hCoV-19/Netherlands/Gelderland_122/2020\|EPI_ISL_46 | ···················· | ···················· | ····················· | ···················· | ···················· | ···················· |
| C | 6137 | hCoV-19/Netherlands/Gelderland_123/2020\|EPI_ISL_46 | ···················· | ···················· | ····················· | ···················· | ···················· | ···················· |
| C | 6138 | hCoV-19/Netherlands/Gelderland_124/2020\|EPI_ISL_46 | ···················· | ···················· | ····················· | ···················· | ···················· | ···················· |
| C | 6139 | hCoV-19/Netherlands/Gelderland_125/2020\|EPI_ISL_46 | ···················· | ···················· | ····················· | ···················· | ···················· | ···················· |
| C | 6140 | hCoV-19/Netherlands/Gelderland_126/2020\|EPI_ISL_46 | ···················· | ···················· | ····················· | ···················· | ···················· | ···················· |
| C | 6141 | hCoV-19/Netherlands/Gelderland_127/2020\|EPI_ISL_46 | ···················· | ···················· | ····················· | ···················· | ···················· | ···················· |
| C | 6142 | hCoV-19/Netherlands/Gelderland_128/2020\|EPI_ISL_46 | ···················· | ···················· | ····················· | ···················· | ···················· | ···················· |
| C | 6143 | hCoV-19/Netherlands/Gelderland_129/2020\|EPI_ISL_46 | ···················· | ···················· | ····················· | ···················· | ···················· | ···················· |
| C | 6144 | hCoV-19/Netherlands/Gelderland_130/2020\|EPI_ISL_46 | ···················· | ···················· | ····················· | ···················· | ···················· | ···················· |
| C | 6145 | hCoV-19/Taiwan/TSGH-33/2020\|EPI_ISL_447592\|2020-03 | ···················· | ···················· | ····················· | ···················· | ···················· | ···················· |
| C | 6146 | hCoV-19/Taiwan/TSGH-34/2020\|EPI_ISL_447593\|2020-04 | ···················· | ···················· | ····················· | ···················· | ···················· | ···················· |
| C | 6147 | hCoV-19/Australia/QLDID941/2020\|EPI_ISL_447594\|202 | ···················· | ···················· | ····················· | ···················· | ···················· | ···················· |
| C | 6148 | hCoV-19/Australia/QLDID942/2020\|EPI_ISL_447595\|202 | ···················· | ···················· | ····················· | ···················· | ···················· | ···················· |
| C | 6149 | hCoV-19/DRC/1151/2020\|EPI_ISL_447596\|2020-04-05 | ···················· | ···················· | ····················· | ···················· | ···················· | ···················· |
| C | 6150 | hCoV-19/DRC/3440/2020\|EPI_ISL_447597\|2020-04-22 | ···················· | ···················· | ····················· | ···················· | ···················· | ···················· |
| C | 6151 | hCoV-19/DRC/3451/2020\|EPI_ISL_447598\|2020-04-22 | ···················· | ···················· | ····················· | ···················· | ···················· | ···················· |
| C | 6152 | hCoV-19/DRC/3453/2020\|EPI_ISL_447599\|2020-04-22 | ···················· | ···················· | ····················· | ···················· | ···················· | ···················· |
| C | 6153 | hCoV-19/England/SHEF-D0300/2020\|EPI_ISL_448902\|202 | ···················· | ···················· | ····················· | ···················· | ···················· | ···················· |

| | | | | | | | | |
|---|---|---|---|---|---|---|---|---|
| C | 6154 | hCoV-19/England/SHEF-D04D0/2020\|EPI_ISL_448903\|202 | .................... | .................... | .................... | .................... | .................... | .................... |
| C | 6155 | hCoV-19/England/SHEF-D0986/2020\|EPI_ISL_448907\|202 | .................... | .................... | .................... | .................... | .................... | .................... |
| C | 6156 | hCoV-19/England/SHEF-D0FB1/2020\|EPI_ISL_448908\|202 | .................... | .................... | .................... | .................... | .................... | .................... |
| C | 6157 | hCoV-19/England/SHEF-D1660/2020\|EPI_ISL_448909\|202 | .................... | .................... | .................... | .................... | .................... | .................... |
| C | 6158 | hCoV-19/England/SHEF-D2720/2020\|EPI_ISL_448913\|202 | .................... | .................... | .................... | .................... | .................... | .................... |
| C | 6161 | hCoV-19/Northern Ireland/NIRE-FAE4B/2020\|EPI_ISL_4 | .................... | .................... | .................... | .................... | .................... | .................... |
| C | 6162 | hCoV-19/Denmark/ALAB-HH-99/2020\|EPI_ISL_452100\|202 | .................... | .................... | .................... | .................... | .................... | .................... |
| C | 6163 | hCoV-19/Denmark/ALAB-SSI-763/2020\|EPI_ISL_452101\|2 | .................... | .................... | .................... | .................... | .................... | .................... |
| C | 6164 | hCoV-19/Denmark/ALAB-SSI-766/2020\|EPI_ISL_452102\|2 | .................... | .................... | .................... | .................... | .................... | .................... |
| C | 6165 | hCoV-19/Germany/BY-MVP-0018/2020\|EPI_ISL_452103\|20 | .................... | .................... | .................... | .................... | .................... | .................... |
| C | 6167 | hCoV-19/USA/TX-CDC-3008/2020\|EPI_ISL_452105\|2020-0 | .................... | .................... | .................... | .................... | .................... | .................... |
| C | 6168 | hCoV-19/USA/LA-CDC-0499/2020\|EPI_ISL_452106\|2020-0 | .................... | .................... | .................... | .................... | .................... | .................... |
| C | 6169 | hCoV-19/USA/LA-CDC-0757/2020\|EPI_ISL_452107\|2020-0 | .................... | .................... | .................... | .................... | .................... | .................... |
| C | 6170 | hCoV-19/USA/MN-CDC-0103/2020\|EPI_ISL_452108\|2020-0 | .................... | .................... | .................... | .................... | .................... | .................... |
| C | 6171 | hCoV-19/USA/NM-CDC-3589/2020\|EPI_ISL_452109\|2020-0 | .................... | .................... | .................... | .................... | .................... | .................... |
| C | 6172 | hCoV-19/USA/FL-CDC-6318/2020\|EPI_ISL_452110\|2020-0 | .................... | .................... | .................... | .................... | .................... | .................... |
| C | 6173 | hCoV-19/USA/FL-CDC-2718/2020\|EPI_ISL_452111\|2020-0 | .................... | .................... | .................... | .................... | .................... | .................... |
| C | 6174 | hCoV-19/USA/TX-CDC-3009/2020\|EPI_ISL_452112\|2020-0 | .................... | .................... | .................... | .................... | .................... | .................... |
| C | 6175 | hCoV-19/USA/MN-CDC-0108/2020\|EPI_ISL_452113\|2020-0 | .................... | .................... | .................... | .................... | .................... | .................... |
| C | 6176 | hCoV-19/USA/KS-CDC-4589/2020\|EPI_ISL_452114\|2020-0 | .................... | .................... | .................... | .................... | .................... | .................... |
| C | 6177 | hCoV-19/USA/TX-CDC-8001/2020\|EPI_ISL_452115\|2020-0 | .................... | .................... | .................... | .................... | .................... | .................... |
| C | 6178 | hCoV-19/USA/NJ-CDC-3036/2020\|EPI_ISL_452116\|2020-0 | .................... | .................... | .................... | .................... | .................... | .................... |
| C | 6179 | hCoV-19/USA/NJ-CDC-7300/2020\|EPI_ISL_452117\|2020-0 | .................... | .................... | .................... | .................... | .................... | .................... |
| C | 6180 | hCoV-19/USA/AZ-CDC-1911/2020\|EPI_ISL_452118\|2020-0 | .................... | .................... | .................... | .................... | .................... | .................... |
| C | 6181 | hCoV-19/USA/VI-CDC-3588/2020\|EPI_ISL_452119\|2020-0 | .................... | .................... | .................... | .................... | .................... | .................... |
| C | 6182 | hCoV-19/USA/GA-CDC-8816/2020\|EPI_ISL_452120\|2020-0 | .................... | .................... | .................... | .................... | .................... | .................... |
| C | 6183 | hCoV-19/USA/VI-CDC-3661/2020\|EPI_ISL_452121\|2020-0 | .................... | .................... | .................... | .................... | .................... | .................... |
| C | 6184 | hCoV-19/USA/VI-CDC-3705/2020\|EPI_ISL_452122\|2020-0 | .................... | .................... | .................... | .................... | .................... | .................... |
| C | 6185 | hCoV-19/USA/VI-CDC-3688/2020\|EPI_ISL_452123\|2020-0 | .................... | .................... | .................... | .................... | .................... | .................... |
| C | 6186 | hCoV-19/USA/NC-CDC-6999/2020\|EPI_ISL_452124\|2020-0 | .................... | .................... | .................... | .................... | .................... | .................... |
| C | 6187 | hCoV-19/USA/GA-CDC-2059/2020\|EPI_ISL_452125\|2020-0 | .................... | .................... | .................... | .................... | .................... | .................... |
| C | 6188 | hCoV-19/USA/NC-CDC-0034/2020\|EPI_ISL_452126\|2020-0 | .................... | .................... | .................... | .................... | .................... | .................... |
| C | 6189 | hCoV-19/USA/CO-CDC-5610/2020\|EPI_ISL_452127\|2020-0 | .................... | .................... | .................... | .................... | .................... | .................... |
| C | 6190 | hCoV-19/USA/CO-CDC-5667/2020\|EPI_ISL_452128\|2020-0 | .................... | .................... | .................... | .................... | .................... | .................... |
| C | 6191 | hCoV-19/USA/CO-CDC-5780/2020\|EPI_ISL_452129\|2020-0 | .................... | .................... | .................... | .................... | .................... | .................... |
| C | 6192 | hCoV-19/USA/CO-CDC-5607/2020\|EPI_ISL_452130\|2020-0 | .................... | .................... | .................... | .................... | .................... | .................... |
| C | 6193 | hCoV-19/USA/IN-CDC-2013/2020\|EPI_ISL_452131\|2020-0 | .................... | .................... | .................... | .................... | .................... | .................... |
| C | 6194 | hCoV-19/USA/FL-CDC-2316/2020\|EPI_ISL_452132\|2020-0 | .................... | .................... | .................... | .................... | .................... | .................... |
| C | 6195 | hCoV-19/USA/MN-CDC-0106/2020\|EPI_ISL_452133\|2020-0 | .................... | .................... | .................... | .................... | .................... | .................... |
| C | 6196 | hCoV-19/USA/IL-CDC-1732/2020\|EPI_ISL_452134\|2020-0 | .................... | .................... | .................... | .................... | .................... | .................... |
| C | 6197 | hCoV-19/USA/VI-CDC-4412/2020\|EPI_ISL_452137\|2020-0 | .................... | .................... | .................... | .................... | .................... | .................... |
| C | 6198 | hCoV-19/USA/VI-CDC-4670/2020\|EPI_ISL_452138\|2020-0 | .................... | .................... | .................... | .................... | .................... | .................... |
| C | 6199 | hCoV-19/Mexico/CMX-InDRE-07/2020\|EPI_ISL_452139\|20 | .................... | .................... | .................... | .................... | .................... | .................... |
| C | 6200 | hCoV-19/Belgium/20A42/2020\|EPI_ISL_452140\|2020-04- | .................... | .................... | .................... | .................... | .................... | .................... |
| C | 6201 | hCoV-19/Belgium/20A37/2020\|EPI_ISL_452142\|2020-03- | .................... | .................... | .................... | .................... | .................... | .................... |
| C | 6202 | hCoV-19/USA/CT-Yale-333/2020\|EPI_ISL_452145\|2020-0 | .................... | .................... | .................... | .................... | .................... | .................... |
| C | 6203 | hCoV-19/Belgium/20A38/2020\|EPI_ISL_452148\|2020-04- | .................... | .................... | .................... | .................... | .................... | .................... |
| C | 6204 | hCoV-19/Belgium/20A44/2020\|EPI_ISL_452149\|2020-04- | .................... | .................... | .................... | .................... | .................... | .................... |
| C | 6205 | hCoV-19/Belgium/20A47/2020\|EPI_ISL_452150\|2020-04- | .................... | .................... | .................... | .................... | .................... | .................... |
| C | 6206 | hCoV-19/Belgium/20A49/2020\|EPI_ISL_452151\|2020-04- | .................... | .................... | .................... | .................... | .................... | .................... |
| C | 6207 | hCoV-19/Belgium/20A55/2020\|EPI_ISL_452152\|2020-04- | .................... | .................... | .................... | .................... | .................... | .................... |
| C | 6208 | hCoV-19/USA/UT-01441/2020\|EPI_ISL_452154\|2020-04-1 | .................... | .................... | .................... | .................... | .................... | .................... |
| C | 6209 | hCoV-19/USA/UT-01450/2020\|EPI_ISL_452157\|2020-04-1 | .................... | .................... | .................... | .................... | .................... | .................... |
| C | 6210 | hCoV-19/USA/UT-01525/2020\|EPI_ISL_452158\|2020-04-1 | .................... | .................... | .................... | .................... | .................... | .................... |
| C | 6211 | hCoV-19/USA/UT-01901/2020\|EPI_ISL_452161\|2020-04-1 | .................... | .................... | .................... | .................... | .................... | .................... |
| C | 6212 | hCoV-19/USA/UT-01943/2020\|EPI_ISL_452163\|2020-04-1 | .................... | .................... | .................... | .................... | .................... | .................... |
| C | 6213 | hCoV-19/USA/UT-01979/2020\|EPI_ISL_452166\|2020-04-1 | .................... | .................... | .................... | .................... | .................... | .................... |
| C | 6214 | hCoV-19/USA/UT-01980/2020\|EPI_ISL_452167\|2020-04-1 | .................... | .................... | .................... | .................... | .................... | .................... |
| C | 6215 | hCoV-19/USA/UT-01981/2020\|EPI_ISL_452168\|2020-04-1 | .................... | .................... | .................... | .................... | .................... | .................... |
| C | 6216 | hCoV-19/USA/UT-01982/2020\|EPI_ISL_452169\|2020-04-1 | .................... | .................... | .................... | .................... | .................... | .................... |
| C | 6217 | hCoV-19/USA/UT-01985/2020\|EPI_ISL_452171\|2020-04-1 | .................... | .................... | .................... | .................... | .................... | .................... |
| C | 6218 | hCoV-19/USA/UT-01986/2020\|EPI_ISL_452172\|2020-04-1 | .................... | .................... | .................... | .................... | .................... | .................... |
| C | 6219 | hCoV-19/USA/UT-01987/2020\|EPI_ISL_452173\|2020-04-1 | .................... | .................... | .................... | .................... | .................... | .................... |
| C | 6220 | hCoV-19/USA/UT-01992/2020\|EPI_ISL_452176\|2020-04-1 | .................... | .................... | .................... | .................... | .................... | .................... |
| C | 6221 | hCoV-19/Taiwan/CGMH-CGU-26/2020\|EPI_ISL_452178\|202 | .................... | .................... | .................... | .................... | .................... | .................... |
| C | 6222 | hCoV-19/Taiwan/CGMH-CGU-27/2020\|EPI_ISL_452179\|202 | .................... | .................... | .................... | .................... | .................... | .................... |
| C | 6223 | hCoV-19/Italy/VEN-IZSVe-10-41/2020\|EPI_ISL_452181\| | .................... | .................... | .................... | .................... | .................... | .................... |
| C | 6224 | hCoV-19/Italy/VEN-IZSVe-11-181/2020\|EPI_ISL_452182 | .................... | .................... | .................... | .................... | .................... | .................... |
| C | 6225 | hCoV-19/Italy/VEN-IZSVe-17-11/2020\|EPI_ISL_452183\| | .................... | .................... | .................... | .................... | .................... | .................... |
| C | 6226 | hCoV-19/Italy/VEN-IZSVe-18-48/2020\|EPI_ISL_452184\| | .................... | .................... | .................... | .................... | .................... | .................... |
| C | 6227 | hCoV-19/Italy/VEN-IZSVe-23-50/2020\|EPI_ISL_452185\| | .................... | .................... | .................... | .................... | .................... | .................... |
| C | 6228 | hCoV-19/Italy/VEN-IZSVe-25-80/2020\|EPI_ISL_452186\| | .................... | .................... | .................... | .................... | .................... | .................... |
| C | 6229 | hCoV-19/Italy/VEN-IZSVe-31-18/2020\|EPI_ISL_452187\| | .................... | .................... | .................... | .................... | .................... | .................... |
| C | 6230 | hCoV-19/Italy/VEN-IZSVe-31-19/2020\|EPI_ISL_452188\| | .................... | .................... | .................... | .................... | .................... | .................... |
| C | 6231 | hCoV-19/Italy/VEN-IZSVe-33-126/2020\|EPI_ISL_452189 | .................... | .................... | .................... | .................... | .................... | .................... |
| C | 6232 | hCoV-19/Italy/VEN-IZSVe-46-8/2020\|EPI_ISL_452190\|2 | .................... | .................... | .................... | .................... | .................... | .................... |
| C | 6233 | hCoV-19/Italy/VEN-IZSVe-46-23/2020\|EPI_ISL_452191\| | .................... | .................... | .................... | .................... | .................... | .................... |
| C | 6234 | hCoV-19/India/MH-NIV-10380/2020\|EPI_ISL_452192\|202 | .................... | .................... | .................... | .................... | .................... | .................... |

| | | | | | | | | | |
|---|---|---|---|---|---|---|---|---|---|
| C | 6235 hCoV-19/India/MH-NIV-10419/2020\|EPI_ISL_452193\|202 | ···················· | ···················· | ···················· | ···················· | ···················· | ···················· | | |
| C | 6237 hCoV-19/India/MH-NIV-11681/2020\|EPI_ISL_452195\|202 | ···················· | ···················· | ···················· | ···················· | ···················· | ···················· | | |
| C | 6238 hCoV-19/India/MH-NIV-11683/2020\|EPI_ISL_452196\|202 | ···················· | ···················· | ···················· | ···················· | ···················· | ···················· | | |
| C | 6239 hCoV-19/India/MH-NIV-11687/2020\|EPI_ISL_452197\|202 | ···················· | ···················· | ···················· | ···················· | ···················· | ···················· | | |
| C | 6240 hCoV-19/India/MH-NIV-11855/2020\|EPI_ISL_452198\|202 | ···················· | ···················· | ···················· | ···················· | ···················· | ···················· | | |
| C | 6241 hCoV-19/India/MH-NIV-12019/2020\|EPI_ISL_452199\|202 | ···················· | ···················· | ···················· | ········------······ | ···················· | ···················· | | 1 |
| C | 6242 hCoV-19/England/LIVE-9F19A/2020\|EPI_ISL_453433\|202 | ···················· | ···················A | ···················· | ···················· | ···················· | ···················· | 1 | |
| C | 6243 hCoV-19/England/LIVE-9F1C7/2020\|EPI_ISL_453435\|202 | ···················· | ···················· | ···················· | ···················· | ···················· | ···················· | | |
| C | 6244 hCoV-19/England/LIVE-9F21F/2020\|EPI_ISL_453437\|202 | ···················· | ···················· | ···················· | ···················· | ···················· | ···················· | | |
| C | 6245 hCoV-19/England/LIVE-9F26A/2020\|EPI_ISL_453441\|202 | ···················· | ···················· | ···················· | ···················· | ···················· | ···················· | | |
| C | 6246 hCoV-19/England/LIVE-9F2D3/2020\|EPI_ISL_453442\|202 | ···········T········ | ···················· | ···················· | ···················· | ···················· | ···················· | | |
| C | 6247 hCoV-19/England/LIVE-9F31C/2020\|EPI_ISL_453446\|202 | ···················· | ···················A | ···················· | ···················· | ···················· | ···················· | 1 | |
| C | 6248 hCoV-19/England/LIVE-9F3B2/2020\|EPI_ISL_453452\|202 | ···················· | ···················· | ···················· | ···················· | ···················· | ···················· | | |
| C | 6249 hCoV-19/England/LIVE-9F3C1/2020\|EPI_ISL_453453\|202 | ···················· | ···················· | ···················· | ···················· | ···················· | ···················· | | |
| C | 6250 hCoV-19/England/LOND-D570F/2020\|EPI_ISL_453460\|202 | ···················· | ···················· | ···················· | ···················· | ···················· | ···················· | | |
| C | 6251 hCoV-19/England/LOND-D5769/2020\|EPI_ISL_453461\|202 | ···················· | ···················· | ···················· | ···················· | ···················· | ···················· | | |
| C | 6252 hCoV-19/England/LOND-D5787/2020\|EPI_ISL_453462\|202 | ···················· | ···················· | ···················· | ···················· | ···················· | ···················· | | |
| C | 6253 hCoV-19/England/LOND-D5FE9/2020\|EPI_ISL_453463\|202 | ···················· | ···················· | ···················· | ···················· | ···················· | ···················· | | |
| C | 6254 hCoV-19/England/LOND-D604F/2020\|EPI_ISL_453464\|202 | ···················· | ···················· | ···················· | ···················· | ···················· | ···················· | | |
| C | 6256 hCoV-19/Northern Ireland/NIRE-101E22/2020\|EPI_ISL_ | ···················· | ···················· | ···················· | ···················· | ···················· | ···················· | | |
| C | 6257 hCoV-19/Northern Ireland/NIRE-101E31/2020\|EPI_ISL_ | ···················· | ···················· | ···················· | ···················· | ···················· | ···················· | | |
| C | 6258 hCoV-19/Northern Ireland/NIRE-101E40/2020\|EPI_ISL_ | ···················· | ···················· | ···················· | ···················· | ···················· | ···················· | | |
| C | 6259 hCoV-19/Northern Ireland/NIRE-101E6E/2020\|EPI_ISL_ | ···················· | ···················· | ···················· | ···················· | ···················· | ···················· | | |
| C | 6260 hCoV-19/Northern Ireland/NIRE-101E7D/2020\|EPI_ISL_ | ···················· | ···················· | ···················· | ···················· | ···················· | ···················· | | |
| C | 6261 hCoV-19/Northern Ireland/NIRE-101E8C/2020\|EPI_ISL_ | ···················· | ···················· | ···················· | ···················· | ···················· | ···················· | | |
| C | 6263 hCoV-19/Northern Ireland/NIRE-101EAA/2020\|EPI_ISL_ | ···················· | ···················· | ···················· | ···················· | ···················· | ···················· | | |
| C | 6266 hCoV-19/Northern Ireland/NIRE-101ED7/2020\|EPI_ISL_ | ···················· | ···················· | ···················· | ···················· | ···················· | ···················· | | |
| C | 6267 hCoV-19/Northern Ireland/NIRE-101EE6/2020\|EPI_ISL_ | ···················· | ···················· | ···················· | ···················· | ···················· | ···················· | | |
| C | 6268 hCoV-19/Northern Ireland/NIRE-101F10/2020\|EPI_ISL_ | ···················· | ···················· | ···················· | ···················· | ···················· | ···················· | | |
| C | 6269 hCoV-19/Northern Ireland/NIRE-101F2F/2020\|EPI_ISL_ | ···················· | ···················· | ···················· | ···················· | ···················· | ···················· | | |
| C | 6270 hCoV-19/Northern Ireland/NIRE-101F4D/2020\|EPI_ISL_ | ···················· | ···················· | ···················· | ···················· | ···················· | ···················· | | |
| C | 6272 hCoV-19/Northern Ireland/NIRE-101FA7/2020\|EPI_ISL_ | ···················· | ···················· | ···················· | ···················· | ···················· | ···················· | | |
| C | 6273 hCoV-19/Northern Ireland/NIRE-101FC5/2020\|EPI_ISL_ | ···················· | ···················· | ···················· | ···················· | ···················· | ···················· | | |
| C | 6274 hCoV-19/Northern Ireland/NIRE-102331/2020\|EPI_ISL_ | ···················· | ···················· | ···················· | ···················· | ···················· | ···················· | | |
| C | 6275 hCoV-19/Northern Ireland/NIRE-102340/2020\|EPI_ISL_ | ···················· | ···················· | ···················· | ···················· | ···················· | ···················· | | |
| C | 6276 hCoV-19/England/NORT-288DF8/2020\|EPI_ISL_453489\|20 | ···················· | ···················· | ···················· | ···················· | ···················· | ···················· | | |
| C | 6277 hCoV-19/England/NORT-288EB9/2020\|EPI_ISL_453492\|20 | ···················· | ···················· | ···················· | ···················· | ···················· | ···················· | | |
| C | 6278 hCoV-19/England/NORT-289243/2020\|EPI_ISL_453499\|20 | ···················· | ···················· | ···················· | ···················· | ···················· | ···················· | | |
| C | 6279 hCoV-19/USA/UT-01677/2020\|EPI_ISL_454707\|2020-04-1 | ···················· | ···················· | ···················· | ···················· | ···················· | ···················· | | |
| C | 6280 hCoV-19/USA/UT-01679/2020\|EPI_ISL_454708\|2020-04-1 | ···················· | ···················· | ···················· | ···················· | ···················· | ···················· | | |
| C | 6281 hCoV-19/USA/UT-01683/2020\|EPI_ISL_454709\|2020-04-1 | ···················· | ···················· | ···················· | ···················· | ···················· | ···················· | | |
| C | 6282 hCoV-19/USA/UT-01686/2020\|EPI_ISL_454710\|2020-04-1 | ···················· | ···················· | ···················· | ···················· | ···················· | ···················· | | |
| C | 6283 hCoV-19/USA/UT-01687/2020\|EPI_ISL_454711\|2020-04-1 | ···················· | ···················· | ···················· | ···················· | ···················· | ···················· | | |
| C | 6284 hCoV-19/USA/UT-01693/2020\|EPI_ISL_454714\|2020-04-1 | ···················· | ···················· | ···················· | ···················· | ···················· | ···················· | | |
| C | 6285 hCoV-19/USA/UT-01694/2020\|EPI_ISL_454715\|2020-04-1 | ···················· | ···················· | ···················· | ···················· | ···················· | ···················· | | |
| C | 6286 hCoV-19/USA/UT-01892/2020\|EPI_ISL_454717\|2020-04-1 | ···················· | ···················· | ···················· | ···················· | ···················· | ···················· | | |
| C | 6287 hCoV-19/USA/UT-01893/2020\|EPI_ISL_454718\|2020-04-1 | ···················· | ···················· | ···················· | ···················· | ···················· | ···················· | | |
| C | 6288 hCoV-19/USA/UT-01896/2020\|EPI_ISL_454721\|2020-04-1 | ···················· | ···················· | ···················· | ···················· | ···················· | ···················· | | |
| C | 6289 hCoV-19/USA/UT-01996/2020\|EPI_ISL_454722\|2020-04-1 | ···················· | ···················· | ···················· | ···················· | ···················· | ···················· | | |
| C | 6290 hCoV-19/USA/UT-01997/2020\|EPI_ISL_454723\|2020-04-1 | ···················· | ···················· | ···················· | ···················· | ···················· | ···················· | | |
| C | 6291 hCoV-19/USA/UT-02004/2020\|EPI_ISL_454725\|2020-04-1 | ···················· | ···················· | ···················· | ···················· | ···················· | ···················· | | |
| C | 6292 hCoV-19/USA/UT-02009/2020\|EPI_ISL_454727\|2020-04-1 | ···················· | ···················· | ···················· | ···················· | ···················· | ···················· | | |
| C | 6293 hCoV-19/USA/UT-03950/2020\|EPI_ISL_454728\|2020-04-1 | ···················· | ···················· | ···················· | ···················· | ···················· | ···················· | | |
| C | 6294 hCoV-19/USA/UT-02016/2020\|EPI_ISL_454729\|2020-04-1 | ···················· | ···················· | ···················· | ···················· | ···················· | ···················· | | |
| C | 6295 hCoV-19/USA/UT-02024/2020\|EPI_ISL_454730\|2020-04-1 | ···················· | ···················· | ···················· | ···················· | ···················· | ···················· | | |
| C | 6296 hCoV-19/USA/UT-02025/2020\|EPI_ISL_454731\|2020-04-1 | ···················· | ···················· | ···················· | ···················· | ···················· | ···················· | | |
| C | 6297 hCoV-19/Russia/Moscow_PMVL-6/2020\|EPI_ISL_454732\|2 | ···················· | ···················· | ···················· | ···················· | ···················· | ···················· | | |
| C | 6298 hCoV-19/Italy/TUS-UniSI-1/2020\|EPI_ISL_454733\|2020 | ···················· | ···················· | ···················· | ···················· | ···················· | ···················· | | |
| C | 6299 hCoV-19/Japan/DP0321/2020\|EPI_ISL_454749\|2020-02-1 | ···················· | ···················· | ···················· | ···················· | ···················· | ···················· | | |
| C | 6300 hCoV-19/Netherlands/NoordHolland_10001/2020\|EPI_IS | ···················· | ···················· | ···················· | ···················· | ···················· | ···················· | | |
| C | 6301 hCoV-19/Netherlands/NoordHolland_10002/2020\|EPI_IS | ···················· | ···················· | ···················· | ···················· | ···················· | ···················· | | |
| C | 6302 hCoV-19/Netherlands/NoordBrabant_10003/2020\|EPI_IS | ···················· | ···················· | ···················· | ···················· | ···················· | ···················· | | |
| C | 6303 hCoV-19/Netherlands/ZuidHolland_10004/2020\|EPI_ISL | ···················· | ···················· | ···················· | ···················· | ···················· | ···················· | | |
| C | 6304 hCoV-19/Netherlands/ZuidHolland_10005/2020\|EPI_ISL | ···················· | ···················· | ···················· | ···················· | ···················· | ···················· | | |
| C | 6305 hCoV-19/Netherlands/NoordHolland_10006/2020\|EPI_IS | ···················· | ···················· | ···················· | ···················· | ···················· | ···················· | | |
| C | 6306 hCoV-19/Netherlands/Utrecht_10007/2020\|EPI_ISL_454 | ···················· | ···················· | ···················· | ···················· | ···················· | ···················· | | |
| C | 6307 hCoV-19/Netherlands/NoordHolland_10008/2020\|EPI_IS | ···················· | ···················· | ···················· | ···················· | ···················· | ···················· | | |
| C | 6308 hCoV-19/Netherlands/Utrecht_10009/2020\|EPI_ISL_454 | ···················· | ···················· | ···················· | ···················· | ···················· | ···················· | | |
| C | 6309 hCoV-19/Netherlands/Utrecht_10010/2020\|EPI_ISL_454 | ···················· | ···················· | ···················· | ···················· | ···················· | ···················· | | |
| C | 6310 hCoV-19/Netherlands/NoordHolland_10011/2020\|EPI_IS | ···················· | ···················· | ···················· | ···················· | ···················· | ···················· | | |
| C | 6311 hCoV-19/Netherlands/Utrecht_10012/2020\|EPI_ISL_454 | ···················· | ···················· | ···················· | ···················· | ···················· | ···················· | | |
| C | 6312 hCoV-19/Netherlands/Utrecht_10013/2020\|EPI_ISL_454 | ···················· | ···················· | ···················· | ···················· | ···················· | ···················· | | |
| C | 6313 hCoV-19/Netherlands/Utrecht_10014/2020\|EPI_ISL_454 | ···················· | ···················· | ···················· | ···················· | ···················· | ···················· | | |
| C | 6314 hCoV-19/Netherlands/Utrecht_10015/2020\|EPI_ISL_454 | ···················· | ···················· | ···················· | ···················· | ···················· | ···················· | | |
| C | 6315 hCoV-19/Netherlands/Utrecht_10016/2020\|EPI_ISL_454 | ···················· | ···················· | ···················· | ···················· | ···················· | ···················· | | |
| C | 6316 hCoV-19/Netherlands/Utrecht_10017/2020\|EPI_ISL_454 | ···················· | ···················· | ···················· | ···················· | ···················· | ···················· | | |
| C | 6317 hCoV-19/Netherlands/Utrecht_10018/2020\|EPI_ISL_454 | ···················· | ···················· | ···················· | ···················· | ···················· | ···················· | | |
| C | 6318 hCoV-19/Netherlands/Utrecht_10019/2020\|EPI_ISL_454 | ···················· | ···················· | ···················· | ···················· | ···················· | ···················· | | |

| | | | | | | | | |
|---|---|---|---|---|---|---|---|---|
| C | 6319 | hCoV-19/Netherlands/Utrecht_10020/2020\|EPI_ISL_454 | ···················· | ···················· | ···················· | ···················· | ···················· | ···················· |
| C | 6320 | hCoV-19/Netherlands/Utrecht_10021/2020\|EPI_ISL_454 | ···················· | ···················· | ···················· | ···················· | ···················· | ···················· |
| C | 6321 | hCoV-19/Netherlands/Utrecht_10022/2020\|EPI_ISL_454 | ···················· | ···················· | ···················· | ···················· | ···················· | ···················· |
| C | 6322 | hCoV-19/Netherlands/Utrecht_10023/2020\|EPI_ISL_454 | ···················· | ···················· | ···················· | ···················· | ···················· | ···················· |
| C | 6323 | hCoV-19/Netherlands/Utrecht_10024/2020\|EPI_ISL_454 | ···················· | ···················· | ···················· | ···················· | ···················· | ···················· |
| C | 6324 | hCoV-19/Netherlands/Utrecht_10025/2020\|EPI_ISL_454 | ···················· | ···················· | ···················· | ···················· | ···················· | ···················· |
| C | 6325 | hCoV-19/Netherlands/Utrecht_10026/2020\|EPI_ISL_454 | ···················· | ···················· | ···················· | ···················· | ···················· | ···················· |
| C | 6326 | hCoV-19/Netherlands/Utrecht_10027/2020\|EPI_ISL_454 | ···················· | ···················· | ···················· | ···················· | ···················· | ···················· |
| C | 6327 | hCoV-19/Netherlands/Utrecht_10028/2020\|EPI_ISL_454 | ···················· | ···················· | ···················· | ···················· | ···················· | ···················· |
| C | 6328 | hCoV-19/Netherlands/Utrecht_10029/2020\|EPI_ISL_454 | ···················· | ···················· | ···················· | ···················· | ···················· | ···················· |
| C | 6329 | hCoV-19/Netherlands/Utrecht_10030/2020\|EPI_ISL_454 | ···················· | ···················· | ···················· | ···················· | ···················· | ···················· |
| C | 6330 | hCoV-19/Netherlands/Utrecht_10031/2020\|EPI_ISL_454 | ···················· | ···················· | ···················· | ···················· | ···················· | ···················· |
| C | 6331 | hCoV-19/Netherlands/Utrecht_10032/2020\|EPI_ISL_454 | ···················· | ···················· | ···················· | ···················· | ···················· | ···················· |
| C | 6332 | hCoV-19/Netherlands/Utrecht_10033/2020\|EPI_ISL_454 | ···················· | ···················· | ···················· | ···················· | ···················· | ···················· |
| C | 6333 | hCoV-19/Netherlands/Utrecht_10034/2020\|EPI_ISL_454 | ···················· | ···················· | ···················· | ···················· | ···················· | ···················· |
| C | 6334 | hCoV-19/Netherlands/Utrecht_10035/2020\|EPI_ISL_454 | ···················· | ···················· | ···················· | ···················· | ···················· | ···················· |
| C | 6335 | hCoV-19/Netherlands/Utrecht_10036/2020\|EPI_ISL_454 | ···················· | ···················· | ···················· | ···················· | ···················· | ···················· |
| C | 6336 | hCoV-19/Netherlands/Utrecht_10037/2020\|EPI_ISL_454 | ···················· | ···················· | ···················· | ···················· | ···················· | ···················· |
| C | 6337 | hCoV-19/Netherlands/Utrecht_10038/2020\|EPI_ISL_454 | ···················· | ···················· | ···················· | ···················· | ···················· | ···················· |
| C | 6338 | hCoV-19/Netherlands/NoordBrabant_10039/2020\|EPI_IS | ···················· | ···················· | ···················· | ···················· | ···················· | ···················· |
| C | 6339 | hCoV-19/Netherlands/NoordBrabant_10040/2020\|EPI_IS | ···················· | ···················· | ···················· | ···················· | ···················· | ···················· |
| C | 6340 | hCoV-19/Netherlands/Utrecht_10041/2020\|EPI_ISL_454 | ···················· | ···················· | ···················· | ···················· | ···················· | ···················· |
| C | 6341 | hCoV-19/Netherlands/NoordBrabant_10042/2020\|EPI_IS | ···················· | ···················· | ···················· | ···················· | ···················· | ···················· |
| C | 6342 | hCoV-19/Netherlands/NoordHolland_10043/2020\|EPI_IS | ···················· | ···················· | ···················· | ···················· | ···················· | ···················· |
| C | 6343 | hCoV-19/Netherlands/Utrecht_10044/2020\|EPI_ISL_454 | ···················· | ···················· | ···················· | ···················· | ···················· | ···················· |
| C | 6344 | hCoV-19/Netherlands/NoordBrabant_10045/2020\|EPI_IS | ···················· | ···················· | ···················· | ···················· | ···················· | ···················· |
| C | 6345 | hCoV-19/Serbia/CA3089-04/2020\|EPI_ISL_454795\|2020- | ···················· | ···················· | ···················· | ···················· | ···················· | ···················· |
| C | 6346 | hCoV-19/USA/NC_200217/2020\|EPI_ISL_454796\|2020-03 | ···················· | ···················· | ···················· | ···················· | ···················· | ···················· |
| C | 6347 | hCoV-19/USA/NC_200219/2020\|EPI_ISL_454797\|2020-03 | ···················· | ···················· | ···················· | ···················· | ···················· | ···················· |
| C | 6348 | hCoV-19/USA/NC_200238/2020\|EPI_ISL_454798\|2020-03 | ···················· | ···················· | ···················· | ···················· | ···················· | ···················· |
| C | 6349 | hCoV-19/USA/NC_200241/2020\|EPI_ISL_454799\|2020-03 | ···················· | ···················· | ···················· | ···················· | ···················· | ···················· |
| C | 6350 | hCoV-19/England/EXET-136194/2020\|EPI_ISL_457206\|20 | ···················· | ···················· | ···················· | ···················· | ···················· | ···················· |
| C | 6351 | hCoV-19/England/EXET-1361A3/2020\|EPI_ISL_457207\|20 | ···················· | ···················· | ···················· | ···················· | ···················· | ···················· |
| C | 6352 | hCoV-19/England/EXET-1361B2/2020\|EPI_ISL_457208\|20 | ···················· | ···················· | ···················· | ···················· | ···················· | ···················· |
| C | 6355 | hCoV-19/England/EXET-136219/2020\|EPI_ISL_457211\|20 | ···················· | ···················· | ···················· | ···················· | ···················· | ···················· |
| C | 6356 | hCoV-19/England/EXET-136246/2020\|EPI_ISL_457212\|20 | ···················· | ···················· | ···················· | ···················· | ···················· | ···················· |
| C | 6358 | hCoV-19/England/EXET-136273/2020\|EPI_ISL_457214\|20 | ···················· | ···················· | ···················· | ···················· | ···················· | ···················· |
| C | 6359 | hCoV-19/USA/CA-SR0368/2020\|EPI_ISL_458251\|2020-04- | ···················· | ···················· | ···················· | ···················· | ···················· | ···················· |
| C | 6360 | hCoV-19/USA/CA-SR0369/2020\|EPI_ISL_458252\|2020-04- | ···················· | ···················· | ···················· | ···················· | ···················· | ···················· |
| C | 6361 | hCoV-19/USA/CA-SR0370/2020\|EPI_ISL_458253\|2020-04- | ···················· | ···················· | ···················· | ···················· | ···················· | ···················· |
| C | 6362 | hCoV-19/USA/CA-SR0371/2020\|EPI_ISL_458254\|2020-04- | ···················· | ·A·················· | ···················· | ···················· | ···················· | ···················· |
| C | 6363 | hCoV-19/USA/CA-SR0372/2020\|EPI_ISL_458255\|2020-04- | ···················· | ···················· | ···················· | ···················· | ···················· | ···················· |
| C | 6364 | hCoV-19/USA/CA-SR0373/2020\|EPI_ISL_458256\|2020-04- | ···················· | ···················· | ···················· | ···················· | ···················· | ···················· |
| C | 6365 | hCoV-19/USA/CA-SR0374/2020\|EPI_ISL_458257\|2020-04- | ···················· | ···················· | ···················· | ···················· | ···················· | ···················· |
| C | 6366 | hCoV-19/USA/CA-SR0376/2020\|EPI_ISL_458258\|2020-04- | ···················· | ···················· | ···················· | ···················· | ···················· | ···················· |
| C | 6367 | hCoV-19/Scotland/GCVR-1702F5/2020\|EPI_ISL_459580\|2 | ···················· | ···················· | ···················· | ···················· | ···················· | ···················· |
| C | 6368 | hCoV-19/Scotland/GCVR-170C42/2020\|EPI_ISL_459581\|2 | ···················· | ···················· | ···················· | ···················· | ···················· | ···················· |
| C | 6369 | hCoV-19/Scotland/GCVR-171397/2020\|EPI_ISL_459583\|2 | ···················· | ···················· | ···················· | ···················· | ···················· | ···················· |
| C | 6370 | hCoV-19/Scotland/GCVR-170CE8/2020\|EPI_ISL_459584\|2 | ···················· | ···················· | ···················· | ···················· | ···················· | ···················· |
| C | 6371 | hCoV-19/Scotland/GCVR-170626/2020\|EPI_ISL_459586\|2 | ···················· | ···················· | ···················· | ···················· | ···················· | ···················· |
| C | 6372 | hCoV-19/Scotland/GCVR-1706AE/2020\|EPI_ISL_459587\|2 | ···················· | ···················· | ···················· | ···················· | ···················· | ···················· |
| C | 6373 | hCoV-19/Scotland/GCVR-17095A/2020\|EPI_ISL_459589\|2 | ···················· | ···················· | ···················· | ···················· | ···················· | ···················· |
| C | 6374 | hCoV-19/Canada/ON_QGLO-04/2020\|EPI_ISL_459868\|2020 | ···················· | ···················· | ···················· | ···················· | ···················· | ···················· |
| C | 6376 | hCoV-19/Canada/ON_QGLO-011/2020\|EPI_ISL_459871\|202 | ···················· | ···················· | ···················· | ···················· | ···················· | ···················· |
| C | 6377 | hCoV-19/Canada/ON_QGLO-012/2020\|EPI_ISL_459872\|202 | ···················· | ···················· | ···················· | ···················· | ···················· | ···················· |
| C | 6378 | hCoV-19/Canada/ON_QGLO-016/2020\|EPI_ISL_459873\|202 | ···················· | ···················· | ···················· | ···················· | ···················· | ···················· |
| C | 6379 | hCoV-19/Canada/ON_QGLO-017/2020\|EPI_ISL_459874\|202 | ···················· | ···················· | ···················· | ···················· | ···················· | ···················· |
| C | 6380 | hCoV-19/Canada/ON_QGLO-018/2020\|EPI_ISL_459875\|202 | ···················· | ···················· | ···················· | ···················· | ···················· | ···················· |
| C | 6381 | hCoV-19/Canada/ON-QGLO-019/2020\|EPI_ISL_459877\|202 | ···················· | ···················· | ···················· | ···················· | ···················· | ···················· |
| C | 6382 | hCoV-19/USA/MI-MDHHS-SC20521/2020\|EPI_ISL_460569\|2 | ···················· | ···················· | ···················· | ···················· | ···················· | ···················· |
| C | 6383 | hCoV-19/USA/MI-MDHHS-SC20523/2020\|EPI_ISL_460571\|2 | ···················· | ···················· | ···················· | ···················· | ···················· | ···················· |
| C | 6384 | hCoV-19/Netherlands/Friesland_35/2020\|EPI_ISL_4608 | ···················· | ···················· | ···················· | ···················· | ···················· | ···················· |
| C | 6385 | hCoV-19/Netherlands/Friesland_36/2020\|EPI_ISL_4608 | ···················· | ···················· | ···················· | ···················· | ···················· | ···················· |
| C | 6386 | hCoV-19/Netherlands/Friesland_37/2020\|EPI_ISL_4608 | ···················· | ···················· | ···················· | ···················· | ···················· | ···················· |
| C | 6387 | hCoV-19/Netherlands/Friesland_38/2020\|EPI_ISL_4608 | ···················· | ···················· | ···················· | ···················· | ···················· | ···················· |
| C | 6388 | hCoV-19/Netherlands/Friesland_39/2020\|EPI_ISL_4608 | ···················· | ···················· | ···················· | ···················· | ···················· | ···················· |
| C | 6389 | hCoV-19/Netherlands/Friesland_40/2020\|EPI_ISL_4608 | ···················· | ···················· | ···················· | ···················· | ···················· | ···················· |
| C | 6390 | hCoV-19/Netherlands/Friesland_41/2020\|EPI_ISL_4608 | ···················· | ···················· | ···················· | ···················· | ···················· | ···················· |
| C | 6391 | hCoV-19/Netherlands/Friesland_44/2020\|EPI_ISL_4608 | ···················· | ···················· | ···················· | ···················· | ······T············· | ···················· |
| C | 6392 | hCoV-19/Netherlands/Gelderland_105/2020\|EPI_ISL_46 | ···················· | ···················· | ···················· | ···················· | ···················· | ···················· |
| C | 6393 | hCoV-19/Netherlands/Gelderland_106/2020\|EPI_ISL_46 | ···················· | ···················· | ···················· | ···················· | ···················· | ···················· |
| C | 6394 | hCoV-19/England/NOTT-1115B1/2020\|EPI_ISL_461894\|20 | ···················· | ···················· | ···················· | ···················· | ···················· | ···················· |
| C | 6395 | hCoV-19/England/NOTT-1115C0/2020\|EPI_ISL_461895\|20 | ···················· | ···················· | ···················· | ···················· | ···················· | ···················· |
| C | 6396 | hCoV-19/England/NOTT-1115DF/2020\|EPI_ISL_461896\|20 | ···················· | ···················· | ···················· | ···················· | ···················· | ···················· |
| C | 6397 | hCoV-19/England/NOTT-1115EE/2020\|EPI_ISL_461897\|20 | ···················· | ···················· | ···················· | ···················· | ···················· | ···················· |
| C | 6398 | hCoV-19/England/NOTT-1115FD/2020\|EPI_ISL_461898\|20 | ···················· | ···················· | ···················· | ···················· | ···················· | ···················· |
| C | 6399 | hCoV-19/England/NOTT-111609/2020\|EPI_ISL_461899\|20 | ···················· | ···················· | ···················· | ···················· | ···················· | ···················· |
| C | 6400 | hCoV-19/Germany/HE-FFM2/2020\|EPI_ISL_529146\|2020-0 | ···················· | ···················· | ···················· | ···················· | ···················· | ···················· |

```
C                 6401 hCoV-19/Spain/VC-FISABIO-569/2020|EPI_ISL_447500|2   ····················  ····················  ·····················  ····················  ····················  ····················
C                 6403 hCoV-19/Spain/VC-FISABIO-571/2020|EPI_ISL_447502|2   ····················  ····················  ·····················  ····················  ····················  ····················
C                 6404 hCoV-19/Spain/VC-FISABIO-572/2020|EPI_ISL_447503|2   ····················  ····················  ·····················  ····················  ····················  ····················
C                 6405 hCoV-19/Spain/VC-FISABIO-575/2020|EPI_ISL_447505|2   ····················  ····················  ·····················  ····················  ····················  ····················
C                 6406 hCoV-19/Spain/VC-FISABIO-576/2020|EPI_ISL_447506|2   ····················  ····················  ·····················  ····················  ····················  ····················
C                 6407 hCoV-19/Spain/VC-FISABIO-580/2020|EPI_ISL_447510|2   ····················  ····················  ·····················  ····················  ····················  ····················
C                 6408 hCoV-19/Spain/VC-FISABIO-581/2020|EPI_ISL_447511|2   ····················  ····················  ·····················  ····················  ····················  ····················
C                 6409 hCoV-19/Spain/VC-FISABIO-582/2020|EPI_ISL_447512|2   ····················  ····················  ·····················  ····················  ····················  ····················
C                 6410 hCoV-19/Spain/VC-FISABIO-583/2020|EPI_ISL_447513|2   ····················  ····················  ·····················  ····················  ····················  ····················
C                 6411 hCoV-19/Spain/VC-FISABIO-584/2020|EPI_ISL_447514|2   ····················  ····················  ·····················  ····················  ····················  ····················
C                 6412 hCoV-19/Spain/VC-FISABIO-585/2020|EPI_ISL_447515|2   ····················  ····················  ·····················  ····················  ····················  ····················
C                 6413 hCoV-19/Spain/VC-FISABIO-594/2020|EPI_ISL_447517|2   ····················  ····················  ·····················  ····················  ····················  ····················
C                 6414 hCoV-19/Spain/VC-FISABIO-597/2020|EPI_ISL_447519|2   ····················  ····················  ·····················  ····················  ····················  ····················
C                 6416 hCoV-19/Spain/VC-FISABIO-603/2020|EPI_ISL_447522|2   ····················  ····················  ·····················  ····················  ····················  ····················
C                 6418 hCoV-19/India/GJ-GBRC87/2020|EPI_ISL_447548|2020-0   ····················  ·A··················  ·····················  ····················  ····················  ··A·················
C                 6419 hCoV-19/India/GJ-GBRC88a/2020|EPI_ISL_447549|2020-   ····················  ····················  ·····················  ····················  ····················  ····················
C                 6420 hCoV-19/India/GJ-GBRC88b/2020|EPI_ISL_447550|2020-   ····················  ····················  ·····················  ····················  ····················  ····················
C                 6421 hCoV-19/India/GJ-GBRC89/2020|EPI_ISL_447551|2020-0   ····················  ····················  ·····················  ····················  ····················  ····················
C                 6422 hCoV-19/India/GJ-GBRC90/2020|EPI_ISL_447552|2020-0   ····················  ····················  ·····················  ····················  ····················  ····················
C                 6423 hCoV-19/India/GJ-GBRC91/2020|EPI_ISL_447553|2020-0   ····················  ·A··················  ·····················  ····················  ····················  ··A·················
C                 6424 hCoV-19/India/GJ-GBRC92/2020|EPI_ISL_447554|2020-0   ····················  ····················  ·····················  ····················  ····················  ····················
C                 6425 hCoV-19/India/GJ-GBRC93/2020|EPI_ISL_447555|2020-0   ····················  ····················  ·····················  ····················  ····················  ····················
C                 6426 hCoV-19/India/TG-CCMB-J021/2020|EPI_ISL_447556|202   ····················  ····················  ·····················  ····················  ····················  ····················
C                 6427 hCoV-19/India/TG-CCMB-J009/2020|EPI_ISL_447557|202   ····················  ····················  ·····················  ····················  ····················  ····················
C                 6428 hCoV-19/India/TG-CCMB-J014/2020|EPI_ISL_447558|202   ····················  ····················  ·····················  ····················  ····················  ····················
C                 6429 hCoV-19/India/TG-CCMB-J068/2020|EPI_ISL_447562|202   ····················  ····················  ·····················  ····················  ····················  ····················
C                 6430 hCoV-19/India/TG-CCMB-J057/2020|EPI_ISL_447563|202   ····················  ····················  ·····················  ····················  ····················  ····················
C                 6431 hCoV-19/India/TG-CCMB-J278/2020|EPI_ISL_447565|202   ····················  ····················  ·····················  ····················  ····················  ····················
C                 6432 hCoV-19/India/TG-CCMB-J131/2020|EPI_ISL_447566|202   ····················  ····················  ·····················  ····················  ····················  ····················
C                 6433 hCoV-19/India/TG-CCMB-J304/2020|EPI_ISL_447567|202   ····················  ····················  ·····················  ····················  ····················  ····················
C                 6434 hCoV-19/India/TG-CCMB-J166/2020|EPI_ISL_447568|202   ····················  ····················  ·····················  ····················  ····················  ····················
C                 6435 hCoV-19/India/TG-CCMB-J206/2020|EPI_ISL_447569|202   ····················  ····················  ·····················  ····················  ····················  ····················
C                 6436 hCoV-19/India/TG-CCMB-J207/2020|EPI_ISL_447570|202   ····················  ····················  ·····················  ····················  ····················  ····················
C                 6437 hCoV-19/India/TG-CCMB-J224/2020|EPI_ISL_447571|202   ····················  ····················  ·····················  ····················  ····················  ····················
C                 6438 hCoV-19/India/TG-CCMB-J230/2020|EPI_ISL_447572|202   ····················  ····················  ·····················  ····················  ····················  ····················
C                 6439 hCoV-19/India/TG-CCMB-J118/2020|EPI_ISL_447573|202   ················C···  ····················  ·····················  ····················  ····················  ····················
C                 6440 hCoV-19/India/TG-CCMB-J208/2020|EPI_ISL_447574|202   ····················  ····················  ·····················  ····················  ····················  ····················
C                 6441 hCoV-19/India/TG-CCMB-J458/2020|EPI_ISL_447575|202   ····················  ····················  ·····················  ····················  ····················  ····················
C                 6442 hCoV-19/India/TG-CCMB-J232/2020|EPI_ISL_447576|202   ····················  ····················  ·····················  ····················  ····················  ····················
C                 6443 hCoV-19/India/TG-CCMB-J321/2020|EPI_ISL_447577|202   ····················  ····················  ·····················  ····················  ····················  ····················
C                 6444 hCoV-19/India/TG-CCMB-J327/2020|EPI_ISL_447578|202   ····················  ····················  ·····················  ····················  ····················  ····················
C                 6445 hCoV-19/India/TG-CCMB-K128/2020|EPI_ISL_447579|202   ····················  ····················  ·····················  ····················  ····················  ····················
C                 6446 hCoV-19/India/TG-CCMB-K496/2020|EPI_ISL_447580|202   ····················  ····················  ·····················  ····················  ····················  ····················
C                 6447 hCoV-19/India/TG-CCMB-K600B/2020|EPI_ISL_447581|20   ····················  ····················  ·····················  ····················  ····················  ····················
C                 6448 hCoV-19/India/TG-CCMB-K601/2020|EPI_ISL_447582|202   ····················  ····················  ·····················  ····················  ····················  ····················
C                 6449 hCoV-19/India/TG-CCMB-L007/2020|EPI_ISL_447583|202   ····················  ····················  ·····················  ····················  ····················  ····················
C                 6450 hCoV-19/India/TN-CCMB-C1-12/2020|EPI_ISL_447584|20   ····················  ····················  ·····················  ····················  ····················  ····················
C                 6451 hCoV-19/India/TN-CCMB-C2-13/2020|EPI_ISL_447585|20   ····················  ····················  ·····················  ····················  ····················  ····················
C                 6452 hCoV-19/India/TN-CCMB-C3-14/2020|EPI_ISL_447586|20   ····················  ····················  ·····················  ····················  ····················  ····················
C                 6453 hCoV-19/India/TN-CCMB-C4-15/2020|EPI_ISL_447587|20   ····················  ····················  ·····················  ····················  ····················  ····················
C                 6454 hCoV-19/USA/FL-UF4/2020|EPI_ISL_447588|2020-03-27    ····················  ····················  ·····················  ····················  ····················  ···············A····
C                 6455 hCoV-19/USA/FL-UF5/2020|EPI_ISL_447589|2020-03-27    ····················  ····················  ·····················  ····················  ····················  ····················
C                 6456 hCoV-19/England/OXON-AF14C/2020|EPI_ISL_448814|202   ····················  ····················  ·····················  ····················  ····················  ····················
C                 6457 hCoV-19/England/OXON-AF179/2020|EPI_ISL_448816|202   ····················  ····················  ·····················  ····················  ····················  ····················
C                 6458 hCoV-19/England/OXON-AF188/2020|EPI_ISL_448817|202   ····················  ····················  ·····················  ····················  ····················  ····················
C                 6459 hCoV-19/England/OXON-AF197/2020|EPI_ISL_448818|202   ····················  ····················  ·····················  ····················  ····················  ····················
C                 6460 hCoV-19/England/OXON-AF249/2020|EPI_ISL_448820|202   ····················  ····················  ·····················  ····················  ····················  ····················
C                 6461 hCoV-19/England/OXON-AF294/2020|EPI_ISL_448821|202   ····················  ···A················  ·····················  ····················  ····················  ····················
C                 6462 hCoV-19/England/OXON-AF2A3/2020|EPI_ISL_448822|202   ····················  ····················  ·····················  ····················  ····················  ····················
C                 6463 hCoV-19/England/OXON-AF319/2020|EPI_ISL_448824|202   ····················  ····················  ·····················  ····················  ····················  ····················
C                 6464 hCoV-19/England/OXON-AF346/2020|EPI_ISL_448825|202   ····················  ····················  ·····················  ····················  ····················  ····················
C                 6465 hCoV-19/England/SHEF-C0CBD/2020|EPI_ISL_448827|202   ····················  ····················  ·····················  ····················  ····················  ····················
C                 6466 hCoV-19/England/SHEF-C0F96/2020|EPI_ISL_448828|202   ····················  ····················  ·····················  ····················  ····················  ····················
C                 6467 hCoV-19/England/SHEF-C1C70/2020|EPI_ISL_448834|202   ····················  ····················  ·····················  ····················  ····················  ····················
C                 6468 hCoV-19/England/SHEF-C1D22/2020|EPI_ISL_448836|202   ····················  ····················  ·····················  ····················  ····················  ····················
C                 6469 hCoV-19/England/SHEF-C2F58/2020|EPI_ISL_448841|202   ····················  ····················  ·····················  ····················  ····················  ····················
C                 6470 hCoV-19/England/SHEF-C41BA/2020|EPI_ISL_448844|202   ····················  ····················  ·····················  ····················  ····················  ····················
C                 6471 hCoV-19/England/SHEF-C4E77/2020|EPI_ISL_448846|202   ····················  ····················  ·····················  ····················  ····················  ····················
C                 6472 hCoV-19/England/SHEF-CBE70/2020|EPI_ISL_448880|202   ····················  ····················  ·····················  ····················  ····················  ····················
C                 6473 hCoV-19/England/SHEF-CC246/2020|EPI_ISL_448881|202   ····················  ····················  ·····················  ····················  ····················  ····················
C                 6474 hCoV-19/England/SHEF-CE1DE/2020|EPI_ISL_448887|202   ····················  ····················  ·····················  ····················  ····················  ····················
C                 6475 hCoV-19/England/SHEF-CF094/2020|EPI_ISL_448894|202   ····················  ····················  ·····················  ····················  ····················  ····················
C                 6476 hCoV-19/England/SHEF-CF86F/2020|EPI_ISL_448896|202   ····················  ····················  ·····················  ····················  ····················  ····················
C                 6477 hCoV-19/Denmark/ALAB-HH-240/2020|EPI_ISL_452080|20   ········T···········  ····················  ···········T·········  ····················  ····················  ····················
C                 6478 hCoV-19/Denmark/ALAB-HH-244/2020|EPI_ISL_452082|20   ····················  ····················  ·····················  ····················  ····················  ····················
C                 6479 hCoV-19/Denmark/ALAB-HH-245/2020|EPI_ISL_452083|20   ····················  ····················  ·····················  ····················  ····················  ····················
C                 6480 hCoV-19/Denmark/ALAB-HH-246/2020|EPI_ISL_452084|20   ····················  ····················  ·····················  ····················  ····················  ····················
C                 6481 hCoV-19/Denmark/ALAB-HH-247/2020|EPI_ISL_452085|20   ····················  ····················  ·····················  ····················  ····················  ····················
```

| | | | | | | | | |
|---|---|---|---|---|---|---|---|---|
| C | 6482 | hCoV-19/Denmark/ALAB-HH-248/2020\|EPI_ISL_452086\|20 | ···················· | ···················· | ···················· | ···················· | ···················· | ···················· |
| C | 6483 | hCoV-19/Denmark/ALAB-HH-249/2020\|EPI_ISL_452087\|20 | ···················· | ···················· | ···················· | ···················· | ···················· | ···················· |
| C | 6484 | hCoV-19/Denmark/ALAB-HH-250/2020\|EPI_ISL_452088\|20 | ···················· | ···················· | ···················· | ···················· | ···················· | ···················· |
| C | 6485 | hCoV-19/Denmark/ALAB-HH-252/2020\|EPI_ISL_452089\|20 | ···················· | ···················· | ···················· | ···················· | ···················· | ···················· |
| C | 6486 | hCoV-19/Denmark/ALAB-HH-254/2020\|EPI_ISL_452090\|20 | ···················· | ···················· | ···················· | ···················· | ···················· | ···················· |
| C | 6487 | hCoV-19/Denmark/ALAB-HH-255/2020\|EPI_ISL_452091\|20 | ···················· | ···················· | ···················· | ···················· | ···················· | ···················· |
| C | 6488 | hCoV-19/Denmark/ALAB-HH-260/2020\|EPI_ISL_452092\|20 | ···················· | ···················· | ···················· | ···················· | ···················· | ···················· |
| C | 6489 | hCoV-19/Denmark/ALAB-HH-262/2020\|EPI_ISL_452093\|20 | ···················· | ···················· | ···················· | ···················· | ···················· | ···················· |
| C | 6490 | hCoV-19/Denmark/ALAB-HH-263/2020\|EPI_ISL_452094\|20 | ···················· | ···················· | ···················· | ···················· | ···················· | ···················· |
| C | 6491 | hCoV-19/Denmark/ALAB-HH-267/2020\|EPI_ISL_452098\|20 | ···················· | ···················· | ···················· | ···················· | ···················· | ···················· |
| C | 6492 | hCoV-19/Denmark/ALAB-HH-97/2020\|EPI_ISL_452099\|202 | ···················· | ···················· | ···················· | ···················· | ···················· | ···················· |
| C | 6493 | hCoV-19/Belgium/Rega-0329237/2020\|EPI_ISL_458207\|2 | ···················· | ···················· | ···················· | ···················· | ···················· | ···················· |
| C | 6494 | hCoV-19/Belgium/Rega-0329238/2020\|EPI_ISL_458208\|2 | ···················· | ···················· | ···················· | ···················· | ···················· | ···················· |
| C | 6495 | hCoV-19/Belgium/Rega-0329239/2020\|EPI_ISL_458209\|2 | ···················· | ···················· | ···················· | ···················· | ···················· | ···················· |
| C | 6496 | hCoV-19/Belgium/Rega-0329240/2020\|EPI_ISL_458210\|2 | ···················· | ···················· | ···················· | ···················· | ···················· | ···················· |
| C | 6497 | hCoV-19/Belgium/Rega-0329241/2020\|EPI_ISL_458211\|2 | ···················· | ···················· | ···················· | ···················· | ···················· | ···················· |
| C | 6498 | hCoV-19/Belgium/Rega-0329242/2020\|EPI_ISL_458212\|2 | ···················· | ···················· | ···················· | ···················· | ···················· | ···················· |
| C | 6499 | hCoV-19/Belgium/Rega-0329243/2020\|EPI_ISL_458213\|2 | ···················· | ···················· | ···················· | ···················· | ···················· | ···················· |
| C | 6500 | hCoV-19/Belgium/Rega-0329244/2020\|EPI_ISL_458214\|2 | ···················· | ···················· | ···················· | ···················· | ···················· | ···················· |
| C | 6501 | hCoV-19/Belgium/Rega-0329245/2020\|EPI_ISL_458215\|2 | ···················· | ···················· | ···················· | ···················· | ···················· | ···················· |
| C | 6502 | hCoV-19/Belgium/Rega-0329246/2020\|EPI_ISL_458216\|2 | ···················· | ···················· | ···················· | ···················· | ···················· | ···················· |
| C | 6503 | hCoV-19/Belgium/Rega-0330247/2020\|EPI_ISL_458217\|2 | ···················· | ···················· | ···················· | ···················· | ···················· | ···················· |
| C | 6504 | hCoV-19/Belgium/Rega-0330248/2020\|EPI_ISL_458218\|2 | ···················· | ···················· | ···················· | ···················· | ···················· | ···················· |
| C | 6505 | hCoV-19/Belgium/Rega-0330249/2020\|EPI_ISL_458219\|2 | ···················· | ···················· | ···················· | ···················· | ···················· | ···················· |
| C | 6506 | hCoV-19/Belgium/Rega-0330250/2020\|EPI_ISL_458220\|2 | ···················· | ···················· | ···················· | ···················· | ···················· | ···················· |
| C | 6507 | hCoV-19/Belgium/Rega-0330251/2020\|EPI_ISL_458221\|2 | ···················· | ···················· | ···················· | ···················· | ···················· | ···················· |
| C | 6508 | hCoV-19/Belgium/Rega-0330252/2020\|EPI_ISL_458222\|2 | ···················· | ···················· | ···················· | ···················· | ···················· | ···················· |
| C | 6509 | hCoV-19/Belgium/Rega-0330253/2020\|EPI_ISL_458223\|2 | ···················· | ···················· | ···················· | ···················· | ···················· | ···················· |
| C | 6510 | hCoV-19/Belgium/Rega-0330254/2020\|EPI_ISL_458224\|2 | ···················· | ···················· | ···················· | ···················· | ···················· | ···················· |
| C | 6511 | hCoV-19/Belgium/Rega-0330255/2020\|EPI_ISL_458225\|2 | ···················· | ···················· | ···················· | ···················· | ···················· | ···················· |
| C | 6512 | hCoV-19/Belgium/Rega-0330256/2020\|EPI_ISL_458226\|2 | ···················· | ···················· | ···················· | ···················· | ···················· | ···················· |
| C | 6513 | hCoV-19/Belgium/Rega-0330257/2020\|EPI_ISL_458227\|2 | ···················· | ···················· | ···················· | ···················· | ···················· | ···················· |
| C | 6514 | hCoV-19/Belgium/Rega-0320258/2020\|EPI_ISL_458228\|2 | ···················· | ···················· | ···················· | ···················· | ···················· | ···················· |
| C | 6515 | hCoV-19/Belgium/Rega-0319259/2020\|EPI_ISL_458229\|2 | ···················· | ···················· | ···················· | ···················· | ···················· | ···················· |
| C | 6516 | hCoV-19/Belgium/Rega-0316260/2020\|EPI_ISL_458230\|2 | ···················· | ···················· | ···················· | ···················· | ···················· | ···················· |
| C | 6517 | hCoV-19/Belgium/Rega-0326261/2020\|EPI_ISL_458231\|2 | ···················· | ···················· | ···················· | ···················· | ···················· | ···················· |
| C | 6518 | hCoV-19/Belgium/Rega-0320262/2020\|EPI_ISL_458232\|2 | ···················· | ···················· | ···················· | ···················· | ···················· | ···················· |
| C | 6519 | hCoV-19/Belgium/Rega-0331263/2020\|EPI_ISL_458233\|2 | ···················· | ···················· | ···················· | ···················· | ···················· | ···················· |
| C | 6520 | hCoV-19/Belgium/Rega-0331264/2020\|EPI_ISL_458234\|2 | ···················· | ···················· | ···················· | ···················· | ···················· | ···················· |
| C | 6521 | hCoV-19/Belgium/Rega-0319265/2020\|EPI_ISL_458235\|2 | ···················· | ···················· | ···················· | ···················· | ···················· | ···················· |
| C | 6522 | hCoV-19/USA/CA-SR0349/2020\|EPI_ISL_458237\|2020-04- | ···················· | ···················· | ···················· | ···················· | ···················· | ···················· |
| C | 6523 | hCoV-19/USA/CA-SR0351/2020\|EPI_ISL_458238\|2020-04- | ···················· | ·A·················· | ···················· | ···················· | ···················· | ···················· |
| C | 6524 | hCoV-19/USA/CA-SR0353/2020\|EPI_ISL_458239\|2020-04- | ···················· | ·A·················· | ···················· | ···················· | ···················· | ···················· |
| C | 6525 | hCoV-19/USA/CA-SR0355/2020\|EPI_ISL_458240\|2020-04- | ···················· | ···················· | ···················· | ···················· | ···················· | ···················· |
| C | 6526 | hCoV-19/USA/CA-SR0356/2020\|EPI_ISL_458241\|2020-04- | ···················· | ···················· | ···················· | ···················· | ···················· | ···················· |
| C | 6527 | hCoV-19/USA/CA-SR0358/2020\|EPI_ISL_458242\|2020-04- | ···················· | ···················· | ···················· | ···················· | ···················· | ···················· |
| C | 6528 | hCoV-19/USA/CA-SR0359/2020\|EPI_ISL_458243\|2020-04- | ···················· | ···················· | ···················· | ···················· | ···················· | ···················· |
| C | 6529 | hCoV-19/USA/CA-SR0360/2020\|EPI_ISL_458244\|2020-04- | ···················· | ·A·················· | ···················· | ···················· | ···················· | ···················· |
| C | 6530 | hCoV-19/USA/CA-SR0361/2020\|EPI_ISL_458245\|2020-04- | ···················· | ···················· | ···················· | ···················· | ···················· | ···················· |
| C | 6531 | hCoV-19/USA/CA-SR0362/2020\|EPI_ISL_458246\|2020-04- | ···················· | ···················· | ···················· | ···················· | ···················· | ···················· |
| C | 6532 | hCoV-19/USA/CA-SR0364/2020\|EPI_ISL_458247\|2020-04- | ···················· | ·A·················· | ···················· | ···················· | ···················· | ···················· |
| C | 6533 | hCoV-19/USA/CA-SR0365/2020\|EPI_ISL_458248\|2020-05- | ···················· | ·A·················· | ···················· | ···················· | ···················· | ···················· |
| C | 6534 | hCoV-19/USA/CA-SR0366/2020\|EPI_ISL_458249\|2020-04- | ···················· | ···················· | ···················· | ···················· | ···················· | ···················· |
| C | 6535 | hCoV-19/USA/CA-SR0367/2020\|EPI_ISL_458250\|2020-04- | ···················· | ···················· | ···················· | ···················· | ···················· | ···················· |
| C | 6536 | hCoV-19/USA/CA-SR0379/2020\|EPI_ISL_458260\|2020-04- | ···················· | ···················· | ···················· | ···················· | ···················· | ···················· |
| C | 6537 | hCoV-19/USA/CA-SR0381/2020\|EPI_ISL_458261\|2020-04- | ···················· | ···················· | ···················· | ···················· | ···················· | ···················· |
| C | 6538 | hCoV-19/USA/CA-SR0382/2020\|EPI_ISL_458262\|2020-04- | ···················· | ···················· | ···················· | ···················· | ···················· | ···················· |
| C | 6539 | hCoV-19/USA/CA-SR0385/2020\|EPI_ISL_458264\|2020-04- | ···················· | ···················· | ···················· | ···················· | ···················· | ···················· |
| C | 6540 | hCoV-19/USA/CA-SR0386/2020\|EPI_ISL_458265\|2020-04- | ···················· | ···················· | ···················· | ···················· | ···················· | ···················· |
| C | 6541 | hCoV-19/USA/CA-SR0387/2020\|EPI_ISL_458266\|2020-04- | ···················· | ···················· | ···················· | ···················· | ···················· | ···················· |
| C | 6542 | hCoV-19/USA/CA-SR0388/2020\|EPI_ISL_458267\|2020-04- | ···················· | ···················· | ···················· | ···················· | ···················· | ···················· |
| C | 6543 | hCoV-19/USA/CA-SR0395/2020\|EPI_ISL_458270\|2020-04- | ···················· | ···················· | ···················· | ···················· | ···················· | ···················· |
| C | 6544 | hCoV-19/USA/CA-SR0398/2020\|EPI_ISL_458271\|2020-04- | ···················· | ·A·················· | ···················· | ···················· | ···················· | ···················· |
| C | 6545 | hCoV-19/USA/CA-SR0399/2020\|EPI_ISL_458272\|2020-04- | ···················· | ·A·················· | ···················· | ···················· | ···················· | ···················· |
| C | 6546 | hCoV-19/USA/CA-SR0402/2020\|EPI_ISL_458274\|2020-04- | ···················· | ···················· | ···················· | ···················· | ···················· | ···················· |
| C | 6547 | hCoV-19/USA/CA-SR0404/2020\|EPI_ISL_458275\|2020-04- | ···················· | ···················· | ···················· | ···················· | ···················· | ···················· |
| C | 6548 | hCoV-19/USA/CA-SR0405/2020\|EPI_ISL_458276\|2020-04- | ···················· | ···················· | ···················· | ···················· | ···················· | ···················· |
| C | 6549 | hCoV-19/USA/CA-SR0407/2020\|EPI_ISL_458277\|2020-04- | ···················· | ···················· | ···················· | ···················· | ···················· | ···················· |
| C | 6550 | hCoV-19/USA/CA-SR0408/2020\|EPI_ISL_458278\|2020-04- | ···················· | ···················· | ···················· | ···················· | ···················· | ···················· |
| C | 6551 | hCoV-19/USA/CA-SR0409/2020\|EPI_ISL_458279\|2020-04- | ···················· | ···················· | ···················· | ···················· | ···················· | ···················· |
| C | 6552 | hCoV-19/USA/CA-SR0410/2020\|EPI_ISL_458280\|2020-04- | ···················· | ···················· | ···················· | ···················· | ···················· | ···················· |
| C | 6553 | hCoV-19/USA/CA-SR0411/2020\|EPI_ISL_458281\|2020-04- | ···················· | ···················· | ···················· | ···················· | ···················· | ···················· |
| C | 6554 | hCoV-19/USA/CA-SR0412/2020\|EPI_ISL_458282\|2020-04- | ···················· | ···················· | ···················· | ···················· | ···················· | ···················· |
| C | 6555 | hCoV-19/USA/CA-SR0413/2020\|EPI_ISL_458283\|2020-03- | ···················· | ···················· | ···················· | ···················· | ···················· | ···················· |
| C | 6556 | hCoV-19/USA/CA-SR0414/2020\|EPI_ISL_458284\|2020-04- | ···················· | ·A·················· | ···················· | ···················· | ···················· | ···················· |
| C | 6557 | hCoV-19/Tunisia/MHT_2/2020\|EPI_ISL_458286\|2020-03- | ···················· | ···················· | ···················· | ···················· | ···················· | ···················· |
| C | 6558 | hCoV-19/Morocco/OUA677_19/2020\|EPI_ISL_458287\|2020 | ··A················· | ···················· | ···················· | ···················· | ···················· | ···················· |
| C | 6559 | hCoV-19/USA/NC_200429/2020\|EPI_ISL_458291\|2020-04 | ···················· | ···················· | ···················· | ···················· | ···················· | ···················· |

| | | | | | | | | | |
|---|---|---|---|---|---|---|---|---|---|
| C | 6560 hCoV-19/USA/NC_200445/2020\|EPI_ISL_458292\|2020-05 | ···················· | ···················· | ···················· | ···················· | ···················· | ···················· | | |
| C | 6561 hCoV-19/USA/NC_200446/2020\|EPI_ISL_458293\|2020-05 | ···················· | ···················· | ···················· | ···················· | ···················· | ···················· | | |
| C | 6562 hCoV-19/USA/NC_200395/2020\|EPI_ISL_458294\|2020-04 | ···················· | ···················· | ···················· | ···················· | ···················· | ···················· | | |
| C | 6563 hCoV-19/USA/NC_200399/2020\|EPI_ISL_458295\|2020-04 | ···················· | ···················· | ···················· | ···················· | ···················· | ···················· | | |
| C | 6564 hCoV-19/USA/NC_200401/2020\|EPI_ISL_458296\|2020-04 | ···················· | ···················· | ···················· | ···················· | ···················· | ···················· | | |
| C | 6565 hCoV-19/USA/NC_200409/2020\|EPI_ISL_458297\|2020-04 | ···················· | ···················· | ···················· | ···················· | ···················· | ···················· | | |
| C | 6566 hCoV-19/India/TG-CCMB-J714/2020\|EPI_ISL_458298\|202 | ···················· | ···················· | ···················· | ···················· | ···················· | ···················· | | |
| C | 6567 hCoV-19/England/CAMB-1ADD1C/2020\|EPI_ISL_459501\|20 | ···················· | ···················· | ···················· | ···················· | ···················· | ···················· | | |
| C | 6568 hCoV-19/England/CAMB-1AE425/2020\|EPI_ISL_459504\|20 | ···················· | ···················· | ···················· | ···················· | ···················· | ···················· | | |
| C | 6569 hCoV-19/Scotland/GCVR-17124F/2020\|EPI_ISL_459538\|2 | ···················· | ···················· | ···················· | ···················· | ···················· | ···················· | | |
| C | 6570 hCoV-19/Scotland/GCVR-17033E/2020\|EPI_ISL_459539\|2 | ···················· | ···················· | ···················· | ···················· | ···················· | ···················· | | |
| C | 6571 hCoV-19/Scotland/GCVR-1710DC/2020\|EPI_ISL_459542\|2 | ···················· | ···················· | ···················· | ···················· | ···················· | ···················· | | |
| C | 6572 hCoV-19/Scotland/GCVR-170C9D/2020\|EPI_ISL_459544\|2 | ···················· | ···················· | ···················· | ···················· | ···················· | ···················· | | |
| C | 6573 hCoV-19/Scotland/GCVR-17131F/2020\|EPI_ISL_459546\|2 | ···················· | ···················A | ···················· | ···················· | ···················· | ···················· | 1 | |
| C | 6574 hCoV-19/Scotland/GCVR-16FFD4/2020\|EPI_ISL_459547\|2 | ···················· | ···················· | ···················· | ···················· | ···················· | ···················· | | |
| C | 6575 hCoV-19/Scotland/GCVR-170459/2020\|EPI_ISL_459548\|2 | ···················· | ···················· | ···················· | ···················· | ···················· | ···················· | | |
| C | 6576 hCoV-19/Scotland/GCVR-170BDC/2020\|EPI_ISL_459550\|2 | ···················· | ···················· | ···················· | ···················· | ···················· | ···················· | | |
| C | 6577 hCoV-19/Scotland/GCVR-170FD0/2020\|EPI_ISL_459552\|2 | ···················· | ···················· | ···················· | ···················· | ···················· | ···················· | | |
| C | 6578 hCoV-19/Scotland/GCVR-170A75/2020\|EPI_ISL_459555\|2 | ···················· | ···················· | ···················· | ···················· | ···················· | ···················· | | |
| C | 6579 hCoV-19/Scotland/GCVR-171388/2020\|EPI_ISL_459556\|2 | ···················· | ···················A | ···················· | ···················· | ···················· | ···················· | 1 | |
| C | 6580 hCoV-19/Scotland/GCVR-1702D7/2020\|EPI_ISL_459557\|2 | ···················· | ·A·················· | ···················· | ···················· | ···················· | ···················· | | |
| C | 6581 hCoV-19/Scotland/GCVR-170116/2020\|EPI_ISL_459558\|2 | ···················· | ·A·················· | ···················· | ···················· | ···················· | ···················· | | |
| C | 6582 hCoV-19/Scotland/GCVR-1704C2/2020\|EPI_ISL_459559\|2 | ···················· | ···················· | ···················· | ···················· | ···················· | ···················· | | |
| C | 6583 hCoV-19/Scotland/GCVR-170082/2020\|EPI_ISL_459560\|2 | ···················· | ···················· | ···················· | ···················· | ···················· | ···················· | | |
| C | 6584 hCoV-19/Scotland/GCVR-170671/2020\|EPI_ISL_459561\|2 | ···················· | ···················· | ···················· | ···················· | ···················· | ···················· | | |
| C | 6585 hCoV-19/Scotland/GCVR-1704FF/2020\|EPI_ISL_459562\|2 | ···················· | ·A·················· | ···················· | ···················· | ···················· | ···················· | | |
| C | 6586 hCoV-19/Scotland/GCVR-17140D/2020\|EPI_ISL_459563\|2 | ···················· | ···················· | ···················· | ···················· | ···················· | ···················· | | |
| C | 6587 hCoV-19/Scotland/GCVR-17117F/2020\|EPI_ISL_459565\|2 | ···················· | ···················· | ···················· | ···················· | ···················· | ···················· | | |
| C | 6588 hCoV-19/Scotland/GCVR-17028C/2020\|EPI_ISL_459566\|2 | ···················· | ···················· | ···················· | ···················· | ···················· | ···················· | | |
| C | 6589 hCoV-19/Scotland/GCVR-1712D6/2020\|EPI_ISL_459568\|2 | ···················· | ·A·················· | ···················· | ···················· | ···················· | ···················· | | |
| C | 6590 hCoV-19/Scotland/GCVR-170B81/2020\|EPI_ISL_459569\|2 | ···················· | ···················· | ···················· | ···················· | ···················· | ···················· | | |
| C | 6591 hCoV-19/Scotland/GCVR-170538/2020\|EPI_ISL_459572\|2 | ···················· | ···················· | ···················· | ···················· | ···················· | ···················· | | |
| C | 6592 hCoV-19/Scotland/GCVR-170073/2020\|EPI_ISL_459574\|2 | ···················· | ···················· | ···················· | ···················· | ···················· | ···················· | | |
| C | 6593 hCoV-19/Scotland/GCVR-170C8E/2020\|EPI_ISL_459575\|2 | ···················· | ···················· | ···················· | ···················· | ···················· | ···················· | | |
| C | 6594 hCoV-19/Scotland/GCVR-171160/2020\|EPI_ISL_459577\|2 | ···················· | ···················· | ···················· | ···················· | ···················· | ···················· | | |
| C | 6595 hCoV-19/Scotland/GCVR-1700DD/2020\|EPI_ISL_459578\|2 | ···················· | ···················A | ···················· | ···················· | ···················· | ···················· | 1 | |
| C | 6596 hCoV-19/Scotland/GCVR-170D12/2020\|EPI_ISL_459590\|2 | ···················· | ···················A | ···················· | ···················· | ···················· | ···················· | 1 | |
| C | 6597 hCoV-19/Scotland/GCVR-170046/2020\|EPI_ISL_459591\|2 | ···················· | ···················· | ···················· | ···················· | ···················· | ···················· | | |
| C | 6598 hCoV-19/Scotland/GCVR-17043B/2020\|EPI_ISL_459593\|2 | ···················· | ···················· | ···················· | ···················· | ···················· | ···················· | | |
| C | 6599 hCoV-19/Scotland/GCVR-170592/2020\|EPI_ISL_459594\|2 | ···················· | ···················· | ···················· | ···················· | ···················· | ···················· | | |
| C | 6600 hCoV-19/Scotland/GCVR-17034D/2020\|EPI_ISL_459595\|2 | ···················· | ···················· | ···················· | ···················· | ···················· | ···················· | | |
| C | 6601 hCoV-19/Scotland/GCVR-1710CD/2020\|EPI_ISL_459596\|2 | ···················· | ···················· | ···················· | ···················· | ···················· | ···················· | | |
| C | 6602 hCoV-19/Scotland/GCVR-1705DE/2020\|EPI_ISL_459597\|2 | ···················· | ···················· | ···················· | ···················· | ···················· | ···················· | | |
| C | 6603 hCoV-19/Scotland/GCVR-170529/2020\|EPI_ISL_459598\|2 | ···················· | ···················· | ···················· | ···················· | ···················· | ···················· | | |
| C | 6604 hCoV-19/USA/MI-MDHHS-SC20508/2020\|EPI_ISL_460556\|2 | ···················· | ···················· | ···················· | ···················· | ···················· | ···················· | | |
| C | 6605 hCoV-19/USA/MI-MDHHS-SC20509/2020\|EPI_ISL_460557\|2 | ···················· | ···················· | ···················· | ···················· | ···················· | ···················· | | |
| C | 6606 hCoV-19/USA/MI-MDHHS-SC20510/2020\|EPI_ISL_460558\|2 | ···················· | ···················· | ···················· | ···················· | ···················· | ···················· | | |
| C | 6607 hCoV-19/USA/MI-MDHHS-SC20511/2020\|EPI_ISL_460559\|2 | ···················· | ···················· | ···················· | ···················· | ···················· | ···················· | | |
| C | 6608 hCoV-19/USA/MI-MDHHS-SC20512/2020\|EPI_ISL_460560\|2 | ···················· | ···················· | ···················· | ···················· | ···················· | ···················· | | |
| C | 6609 hCoV-19/USA/MI-MDHHS-SC20524/2020\|EPI_ISL_460572\|2 | ···················· | ···················· | ···················· | ···················· | ···················· | ···················· | | |
| C | 6610 hCoV-19/USA/MI-MDHHS-SC20525/2020\|EPI_ISL_460573\|2 | ···················· | ···················· | ···················· | ···················· | ···················· | ···················· | | |
| C | 6611 hCoV-19/USA/MI-MDHHS-SC20526/2020\|EPI_ISL_460574\|2 | ···················· | ···················· | ···················· | ···················· | ···················· | ···················· | | |
| C | 6612 hCoV-19/USA/MI-MDHHS-SC20533/2020\|EPI_ISL_460581\|2 | ···················· | ···················· | ···················· | ···················· | ···················· | ···················· | | |
| C | 6613 hCoV-19/USA/MI-MDHHS-SC20534/2020\|EPI_ISL_460582\|2 | ···················· | ···················· | ···················· | ···················· | ···················· | ···················· | | |
| C | 6614 hCoV-19/USA/MI-MDHHS-SC20535/2020\|EPI_ISL_460583\|2 | ···················· | ···················· | ···················· | ···················· | ···················· | ···················· | | |
| C | 6615 hCoV-19/USA/MI-MDHHS-SC20536/2020\|EPI_ISL_460584\|2 | ···················· | ···················· | ···················· | ···················· | ···················· | ···················· | | |
| C | 6616 hCoV-19/USA/MI-MDHHS-SC20537/2020\|EPI_ISL_460585\|2 | ···················· | ···················· | ···················· | ···················· | ···················· | ···················· | | |
| C | 6617 hCoV-19/USA/MI-MDHHS-SC20538/2020\|EPI_ISL_460586\|2 | ···················· | ···················· | ···················· | ···················· | ···················· | ···················· | | |
| C | 6618 hCoV-19/USA/MI-MDHHS-SC20539/2020\|EPI_ISL_460587\|2 | ···················· | ···················· | ···················· | ···················· | ···················· | ···················· | | |
| C | 6619 hCoV-19/USA/MI-MDHHS-SC20540/2020\|EPI_ISL_460588\|2 | ···················· | ···················· | ···················· | ···················· | ···················· | ···················· | | |
| C | 6620 hCoV-19/USA/MI-MDHHS-SC20551/2020\|EPI_ISL_460599\|2 | ···················· | ···················· | ···················· | ···················· | ···················· | ···················· | | |
| C | 6621 hCoV-19/England/NORT-28C170/2020\|EPI_ISL_461801\|20 | ···················· | ···················· | ···················· | ···················· | ···················· | ···················· | | |
| C | 6624 hCoV-19/England/NORW-E8DCB/2020\|EPI_ISL_461809\|202 | ···················· | ···················· | ···················· | ···················· | NNN················· | ···················· | | 1 |
| C | 6629 hCoV-19/England/NORW-E8F2F/2020\|EPI_ISL_461814\|202 | ···················· | ···················· | ···················· | ···················· | ···················· | ···················· | | |
| C | 6632 hCoV-19/England/NORW-E9CBE/2020\|EPI_ISL_461850\|202 | ···················· | ···················· | ···················· | ···················· | ···················· | ···················· | | |
| C | 6652 hCoV-19/England/NORW-EAB65/2020\|EPI_ISL_461884\|202 | ···················· | ···················· | ···················· | ···················· | ···················· | ···················· | | |
| C | 6657 hCoV-19/Spain/VC-FISABIO-609/2020\|EPI_ISL_447527\|2 | ···················· | ···················· | ···················· | ···················· | ···················· | ···················· | | |
| C | 6659 hCoV-19/Spain/VC-FISABIO-611/2020\|EPI_ISL_447529\|2 | ···················· | ···················· | ···················· | ···················· | ···················· | ···················· | | |
| C | 6661 hCoV-19/Spain/VC-FISABIO-613/2020\|EPI_ISL_447531\|2 | ···················· | ···················· | ···················· | ···················· | ···················· | ···················· | | |
| C | 6662 hCoV-19/Spain/CT-HUVH-VH1449/2020\|EPI_ISL_447532\|2 | ···················· | ···················· | ···················· | ···················· | ···················· | ···················· | | |
| C | 6663 hCoV-19/Spain/CT-HUVH-VH7663/2020\|EPI_ISL_447533\|2 | ···················· | ···················· | ···················· | ···················· | ···················· | ···················· | | |
| C | 6664 hCoV-19/India/GJ-GBRC76/2020\|EPI_ISL_447534\|2020-0 | ···················· | ···················· | ···················· | ···················· | ···················· | ···················· | | |
| C | 6665 hCoV-19/India/GJ-GBRC77/2020\|EPI_ISL_447535\|2020-0 | ···················· | ···················· | ···················· | ···················· | ···················· | ···················· | | |
| C | 6666 hCoV-19/India/GJ-GBRC78a/2020\|EPI_ISL_447536\|2020- | ···················· | ···················· | ···················· | ···················· | ···················· | ···················· | | |
| C | 6667 hCoV-19/India/GJ-GBRC78b/2020\|EPI_ISL_447537\|2020- | ···················· | ···················· | ···················· | ···················· | ···················· | ···················· | | |
| C | 6668 hCoV-19/India/GJ-GBRC79a/2020\|EPI_ISL_447538\|2020- | ···················· | ···················· | ···················· | ···················· | ···················· | ···················· | | |
| C | 6669 hCoV-19/India/GJ-GBRC79b/2020\|EPI_ISL_447539\|2020- | ···················· | ···················· | ···················· | ···················· | ···················· | ···················· | | |
| C | 6670 hCoV-19/India/GJ-GBRC81a/2020\|EPI_ISL_447540\|2020- | ···················· | ···················· | ···················· | ···················· | ···················· | ···················· | | |

| | | | | | | | | |
|---|---|---|---|---|---|---|---|---|
| C | 6671 hCoV-19/India/GJ-GBRC81b/2020\|EPI_ISL_447541\|2020- | .................... | .................... | .................... | .................... | .................... | .................... | |
| C | 6672 hCoV-19/India/GJ-GBRC82a/2020\|EPI_ISL_447542\|2020- | .................... | .................... | .................... | .................... | .................... | .................... | |
| C | 6673 hCoV-19/India/GJ-GBRC82b/2020\|EPI_ISL_447543\|2020- | .................... | .................... | .................... | .................... | .................... | .................... | |
| C | 6674 hCoV-19/India/GJ-GBRC83/2020\|EPI_ISL_447544\|2020-0 | .................... | .................... | .................... | .................... | .................... | .................... | |
| C | 6675 hCoV-19/India/GJ-GBRC84/2020\|EPI_ISL_447545\|2020-0 | .................... | .................... | .................... | .................... | .................... | .................... | |
| C | 6676 hCoV-19/India/GJ-GBRC85/2020\|EPI_ISL_447546\|2020-0 | .................... | .A.................. | .................... | .................... | .................... | ...A................ | |
| C | 6677 hCoV-19/India/GJ-GBRC86/2020\|EPI_ISL_447547\|2020-0 | .................... | .................... | .................... | .................... | .................... | .................... | |
| C | 6678 hCoV-19/India/TG-CCMB-J048/2020\|EPI_ISL_447559\|202 | .................... | .................... | .................... | .................... | .................... | .................... | |
| C | 6679 hCoV-19/India/TG-CCMB-J043/2020\|EPI_ISL_447560\|202 | .................... | .................... | .................... | .................... | .................... | .................... | |
| C | 6680 hCoV-19/India/TG-CCMB-J067/2020\|EPI_ISL_447561\|202 | .................... | .................... | .................... | .................... | .................... | .................... | |
| C | 6681 hCoV-19/India/TG-CCMB-J212/2020\|EPI_ISL_447564\|202 | .................... | .................... | .................... | .................... | .................... | .................... | |
| C | 6682 hCoV-19/England/SHEF-C6B7E/2020\|EPI_ISL_448860\|202 | .................... | .................... | .................... | .................... | .................... | .................... | |
| C | 6683 hCoV-19/England/SHEF-C7B31/2020\|EPI_ISL_448864\|202 | .................... | .................... | .................... | .................... | .................... | .................... | |
| C | 6684 hCoV-19/England/SHEF-C81F2/2020\|EPI_ISL_448865\|202 | .................... | .................... | .................... | .................... | .................... | .................... | |
| C | 6685 hCoV-19/England/SHEF-C8286/2020\|EPI_ISL_448866\|202 | .................... | .................... | .................... | .................... | .................... | .................... | |
| C | 6686 hCoV-19/England/SHEF-C82D1/2020\|EPI_ISL_448867\|202 | .................... | .................... | .................... | .................... | .................... | .................... | |
| C | 6687 hCoV-19/England/SHEF-C8DD0/2020\|EPI_ISL_448869\|202 | .................... | .................... | .................... | .................... | .................... | .................... | |
| C | 6688 hCoV-19/England/SHEF-C9100/2020\|EPI_ISL_448870\|202 | .................... | .................... | .................... | .................... | .................... | .................... | |
| C | 6689 hCoV-19/England/SHEF-CA3BE/2020\|EPI_ISL_448874\|202 | .................... | .................... | .................... | .................... | .................... | .................... | |
| C | 6690 hCoV-19/England/SHEF-CB62D/2020\|EPI_ISL_448877\|202 | .................... | .................... | .................... | .................... | .................... | .................... | |
| C | 6691 hCoV-19/India/MH-NIV-12067/2020\|EPI_ISL_452201\|202 | .................... | .................... | .................... | .................... | .................... | .................... | |
| C | 6692 hCoV-19/India/MH-NIV-2810/2020\|EPI_ISL_452202\|2020 | .................... | .................... | .................... | .................... | .................... | .................... | |
| C | 6693 hCoV-19/India/MH-NIV-4367/2020\|EPI_ISL_452203\|2020 | .................... | .................... | .................... | .................... | .................... | .................... | |
| C | 6694 hCoV-19/India/MH-NIV-5109/2020\|EPI_ISL_452204\|2020 | .................... | .................... | .................... | .........------..... | .................... | .................... | 1 |
| C | 6695 hCoV-19/India/MH-NIV-5119/2020\|EPI_ISL_452205\|2020 | .................... | .................... | .................... | .................... | .................... | .................... | |
| C | 6697 hCoV-19/India/MH-NIV-7439/2020\|EPI_ISL_452207\|2020 | .................... | .................... | .................... | .................... | .................... | .................... | |
| C | 6698 hCoV-19/India/MH-NIV-7786/2020\|EPI_ISL_452209\|2020 | .................... | .................... | .................... | .................... | .................... | .................... | |
| C | 6699 hCoV-19/India/MH-NIV-9768/2020\|EPI_ISL_452212\|2020 | .................... | .................... | .................... | .................... | .................... | .................... | |
| C | 6700 hCoV-19/India/MH-NIV-981/2020\|EPI_ISL_452213\|2020- | .................... | .................... | .................... | .................... | .................... | .................... | |
| C | 6701 hCoV-19/India/MH-NIV-QA-709/2020\|EPI_ISL_452214\|20 | .................... | .................... | .................... | .................... | .................... | .................... | |
| C | 6702 hCoV-19/India/MH-NIV-QC-797/2020\|EPI_ISL_452215\|20 | .................... | .................... | .................... | .................... | .................... | .................... | |
| C | 6703 hCoV-19/India/MH-NIV-QC-802/2020\|EPI_ISL_452216\|20 | .................... | .................... | .................... | .................... | .................... | .................... | |
| C | 6704 hCoV-19/India/MH-NIV-QC-800/2020\|EPI_ISL_452217\|20 | .................... | .................... | .................... | .................... | .................... | .................... | |
| C | 6705 hCoV-19/Germany/HE-FFM1/2020\|EPI_ISL_452218\|2020-0 | .................... | .A.................. | .................... | .................... | .................... | .................... | |
| C | 6706 hCoV-19/Germany/HE-FFM3/2020\|EPI_ISL_452219\|2020-0 | .................... | .................... | .................... | .................... | .................... | .................... | |
| C | 6707 hCoV-19/Germany/HE-FFM4/2020\|EPI_ISL_452220\|2020-0 | .................... | .................... | .................... | .................... | .................... | .................... | |
| C | 6708 hCoV-19/Germany/HE-FFM5/2020\|EPI_ISL_452221\|2020-0 | .................... | .................... | .................... | .................... | .................... | .................... | |
| C | 6709 hCoV-19/Germany/HE-FFM6/2020\|EPI_ISL_452222\|2020-0 | .................... | .................... | .................... | .................... | .................... | .................... | |
| C | 6710 hCoV-19/Germany/HE-FFM7/2020\|EPI_ISL_452223\|2020-0 | .................... | .................... | .................... | .................... | .................... | .................... | |
| C | 6711 hCoV-19/Sweden/20-05819/2020\|EPI_ISL_452229\|2020-0 | .................... | .................... | .................... | .................... | .................... | .................... | |
| C | 6712 hCoV-19/Sweden/20-06165/2020\|EPI_ISL_452230\|2020-0 | .................... | .................... | .................... | .................... | .................... | .................... | |
| C | 6713 hCoV-19/Sweden/20-06167/2020\|EPI_ISL_452231\|2020-0 | .................... | .................... | .................... | .................... | .................... | .................... | |
| C | 6714 hCoV-19/Sweden/20-06169/2020\|EPI_ISL_452232\|2020-0 | .................... | .................... | .................... | .................... | .................... | .................... | |
| C | 6715 hCoV-19/Sweden/20-06209/2020\|EPI_ISL_452233\|2020-0 | .................... | .................... | .................... | .................... | .................... | .................... | |
| C | 6716 hCoV-19/Sweden/20-07156/2020\|EPI_ISL_452234\|2020-0 | .................... | .................... | .................... | .................... | .................... | .................... | |
| C | 6717 hCoV-19/Sweden/20-07295/2020\|EPI_ISL_452235\|2020-0 | .................... | .................... | .................... | .................... | .................... | .................... | |
| C | 6718 hCoV-19/Sweden/20-07394/2020\|EPI_ISL_452236\|2020-0 | .................... | .................... | .................... | .................... | .................... | .................... | |
| C | 6719 hCoV-19/Sweden/20-07856/2020\|EPI_ISL_452237\|2020-0 | .................... | .................... | .................... | .................... | .................... | .................... | |
| C | 6720 hCoV-19/Sweden/20-07908/2020\|EPI_ISL_452238\|2020-0 | .................... | .................... | .................... | .................... | .................... | .................... | |
| C | 6721 hCoV-19/Sweden/20-07909/2020\|EPI_ISL_452239\|2020-0 | .................... | .................... | .................... | .................... | .................... | .................... | |
| C | 6722 hCoV-19/Sweden/20-07921/2020\|EPI_ISL_452240\|2020-0 | .................... | .................... | .................... | .................... | .................... | .................... | |
| C | 6723 hCoV-19/Sweden/20-14014/2020\|EPI_ISL_452241\|2020-0 | .................... | .................... | .................... | .................... | .................... | .................... | |
| C | 6724 hCoV-19/Sweden/20-51160/2020\|EPI_ISL_452242\|2020-0 | .................... | .................... | .................... | .................... | .................... | .................... | |
| C | 6725 hCoV-19/England/201060006/2020\|EPI_ISL_452251\|2020 | .................... | .................... | .................... | .................... | .................... | ....N............... | |
| C | 6726 hCoV-19/England/201061090/2020\|EPI_ISL_452252\|2020 | .................... | .................... | .................... | .................... | .................... | .................... | |
| C | 6727 hCoV-19/England/20109003304/2020\|EPI_ISL_452254\|20 | .................... | .................... | .................... | .................... | .................... | .................... | |
| C | 6729 hCoV-19/England/20109045704/2020\|EPI_ISL_452257\|20 | .................... | .................... | .................... | .................... | .................... | .................... | |
| C | 6730 hCoV-19/USA/MI-MDHHS-SC20265/2020\|EPI_ISL_452262\|2 | .................... | .................... | .................... | .................... | .................... | .................... | |
| C | 6731 hCoV-19/USA/MI-MDHHS-SC20390/2020\|EPI_ISL_452264\|2 | .................... | .................... | .................... | .................... | .................... | .................... | |
| C | 6732 hCoV-19/USA/MI-MDHHS-SC20392/2020\|EPI_ISL_452266\|2 | .................... | .................... | .................... | .................... | .................... | .................... | |
| C | 6733 hCoV-19/USA/MI-MDHHS-SC20396/2020\|EPI_ISL_452270\|2 | .................... | .................... | .................... | .................... | .................... | .................... | |
| C | 6734 hCoV-19/USA/MI-MDHHS-SC20398/2020\|EPI_ISL_452272\|2 | .................... | .................... | .................... | .................... | .................... | .................... | |
| C | 6735 hCoV-19/USA/MI-MDHHS-SC20400/2020\|EPI_ISL_452274\|2 | .................... | .................... | .................... | .................... | .................... | .................... | |
| C | 6736 hCoV-19/USA/MI-MDHHS-SC20404/2020\|EPI_ISL_452278\|2 | .................... | .................... | .................... | .................... | .................... | .................... | |
| C | 6737 hCoV-19/USA/MI-MDHHS-SC20405/2020\|EPI_ISL_452279\|2 | .................... | .................... | .................... | .................... | .................... | .................... | |
| C | 6738 hCoV-19/USA/MI-MDHHS-SC20406/2020\|EPI_ISL_452280\|2 | .................... | .................... | .................... | .................... | .................... | .................... | |
| C | 6739 hCoV-19/USA/MI-MDHHS-SC20407/2020\|EPI_ISL_452281\|2 | .................... | .................... | .................... | .................... | .................... | .................... | |
| C | 6740 hCoV-19/USA/MI-MDHHS-SC20408/2020\|EPI_ISL_452282\|2 | .................... | .................... | .................... | .................... | .................... | .................... | |
| C | 6741 hCoV-19/USA/MI-MDHHS-SC20409/2020\|EPI_ISL_452283\|2 | .................... | .................... | .................... | .................... | .................... | .................... | |
| C | 6742 hCoV-19/USA/MI-MDHHS-SC20410/2020\|EPI_ISL_452284\|2 | .................... | .................... | .................... | .................... | .................... | .................... | |
| C | 6743 hCoV-19/USA/MI-MDHHS-SC20411/2020\|EPI_ISL_452285\|2 | .................... | .................... | .................... | .................... | .................... | .................... | |
| C | 6744 hCoV-19/USA/MI-MDHHS-SC20413/2020\|EPI_ISL_452287\|2 | .................... | .................... | .................... | .................... | .................... | .................... | |
| C | 6745 hCoV-19/USA/MI-MDHHS-SC20414/2020\|EPI_ISL_452288\|2 | .................... | .................... | .................... | .................... | .................... | .................... | |
| C | 6746 hCoV-19/USA/MI-MDHHS-SC20415/2020\|EPI_ISL_452289\|2 | .................... | .................... | .................... | .................... | .................... | .................... | |
| C | 6747 hCoV-19/USA/MI-MDHHS-SC20416/2020\|EPI_ISL_452290\|2 | .................... | .................... | .................... | .................... | .................... | .................... | |
| C | 6748 hCoV-19/USA/MI-MDHHS-SC20417/2020\|EPI_ISL_452291\|2 | .................... | .................... | .................... | .................... | .................... | .................... | |
| C | 6749 hCoV-19/USA/MI-MDHHS-SC20418/2020\|EPI_ISL_452292\|2 | .................... | .................... | .................... | .................... | .................... | .................... | |
| C | 6750 hCoV-19/USA/MI-MDHHS-SC20420/2020\|EPI_ISL_452294\|2 | .................... | .................... | .................... | .................... | .................... | .................... | |

C 6751 hCoV-19/USA/MI-MDHHS-SC20421/2020|EPI_ISL_452295|2 ···················· ···················· ···················· ···················· ···················· ····················
C 6752 hCoV-19/USA/MI-MDHHS-SC20423/2020|EPI_ISL_452297|2 ···················· ···················· ···················· ···················· ···················· ····················
C 6753 hCoV-19/USA/MI-MDHHS-SC20424/2020|EPI_ISL_452298|2 ···················· ···················· ···················· ···················· ···················· ····················
C 6754 hCoV-19/England/NORT-28994E/2020|EPI_ISL_453507|20 ···················· ···················· ···················· ···················· ···················· ····················
C 6755 hCoV-19/England/NORT-2899D5/2020|EPI_ISL_453509|20 ···················· ···················· ···················· ···················· ···················· ····················
C 6756 hCoV-19/England/NORT-290D39/2020|EPI_ISL_453534|20 ···················· ···················· ···················· ···················· ···················· ····················
C 6757 hCoV-19/England/NORT-2910B7/2020|EPI_ISL_453538|20 ···················· ···················· ···················· ···················· ···················· ····················
C 6758 hCoV-19/England/NORT-291AD7/2020|EPI_ISL_453544|20 ···················· ···················· ···················· ···················· ···················· ····················
C 6759 hCoV-19/England/NORW-E6DAF/2020|EPI_ISL_453545|202 ···················· ···················· ···················· ···················· ···················· ····················
C 6760 hCoV-19/England/NORW-E6E33/2020|EPI_ISL_453546|202 ···················· ···················· ···················· ···················· ···················· ····················
C 6761 hCoV-19/England/NORW-E6F7C/2020|EPI_ISL_453547|202 ···················· ···················· ···················· ···················· ···················· ····················
C 6762 hCoV-19/England/NORW-E6FD6/2020|EPI_ISL_453548|202 ···················· ···················· ···················· ···················· ···················· ····················
C 6763 hCoV-19/England/NORW-E704B/2020|EPI_ISL_453550|202 ···················· ···················· ···················· ···················· ···················· ····················
C 6764 hCoV-19/England/NORW-E70C3/2020|EPI_ISL_453551|202 ···················· ···················· ···················· ···················· ···················· ····················
C 6765 hCoV-19/England/NORW-E710C/2020|EPI_ISL_453552|202 ···················· ···················· ···················· ···················· ···················· ····················
C 6766 hCoV-19/England/NORW-E7157/2020|EPI_ISL_453553|202 ···················· ···················· ···················· ···················· ···················· ····················
C 6767 hCoV-19/England/NORW-E71EE/2020|EPI_ISL_453554|202 ···················· ···················· ···················· ···················· ···················· ····················
C 6768 hCoV-19/England/NORW-E7236/2020|EPI_ISL_453555|202 ···················· ···················· ···················· ···················· ···················· ····················
C 6769 hCoV-19/England/NORW-E72EB/2020|EPI_ISL_453556|202 ···················· ···················· ···················· ···················· ···················· ····················
C 6770 hCoV-19/England/NORW-E8DF8/2020|EPI_ISL_453558|202 ···················· ···················· ···················· ···················· ···················· ····················
C 6771 hCoV-19/England/NORW-E8E13/2020|EPI_ISL_453559|202 ···················· ···················· ··················Y· ···················· ···················· ···················· 1
C 6772 hCoV-19/England/NORW-E8E31/2020|EPI_ISL_453560|202 ···················· ···················· ···················· ···················· ···················· ····················
C 6773 hCoV-19/England/NORW-E8EF5/2020|EPI_ISL_453563|202 ···················· ···················· ···················· ···················· ···················· ····················
C 6774 hCoV-19/England/NORW-E8F3E/2020|EPI_ISL_453565|202 ···················· ···················· ···················· ···················· ···················· ····················
C 6776 hCoV-19/England/NORW-E8F4D/2020|EPI_ISL_453566|202 ···················· ···················· ···················· ···················· ···················· ····················
C 6777 hCoV-19/England/NORW-E8F5C/2020|EPI_ISL_453567|202 ···················· ···················· ···················· ···················· ···················· ····················
C 6778 hCoV-19/England/NORW-E8F7A/2020|EPI_ISL_453569|202 ···················· ···················· ···················· ···················· ···················· ····················
C 6779 hCoV-19/England/NORW-E8F89/2020|EPI_ISL_453570|202 ···················· ···················· ···················· ···················· NNN················· NNN················· 1
C 6780 hCoV-19/England/NORW-E8FB6/2020|EPI_ISL_453571|202 ···················· ···················· ···················· ···················· NNN················· ···················· 1
C 6781 hCoV-19/England/NORW-E8FD4/2020|EPI_ISL_453572|202 ···················· ···················· ···················· ···················· ···················· ····················
C 6782 hCoV-19/England/NORW-E900D/2020|EPI_ISL_453573|202 ···················· ···················· ···················· ···················· ···················· ····················
C 6783 hCoV-19/England/NORW-E901C/2020|EPI_ISL_453574|202 ···················· ···················· ···················· ···················· ···················· ····················
C 6784 hCoV-19/England/NORW-E902B/2020|EPI_ISL_453575|202 ···················· ···················· ···················· ···················· ···················· ····················
C 6785 hCoV-19/England/NORW-E9049/2020|EPI_ISL_453577|202 ···················· ···················· ···················· ···················· ···················· ····················
C 6786 hCoV-19/England/NORW-E9067/2020|EPI_ISL_453579|202 ···················· ···················· ···················· ···················· ···················· ····················
C 6787 hCoV-19/England/NORW-E9085/2020|EPI_ISL_453580|202 ···················· ···················· ···················· ···················· ···················· ····················
C 6788 hCoV-19/England/NORW-E91CE/2020|EPI_ISL_453582|202 ···················· ···················· ···················· ···················· ···················· ····················
C 6789 hCoV-19/England/NORW-E9216/2020|EPI_ISL_453583|202 ···················· ···················· ···················· ···················· ···················· ····················
C 6790 hCoV-19/England/NORW-E9243/2020|EPI_ISL_453584|202 ···················· ···················· ···················· ···················· ···················· ····················
C 6791 hCoV-19/England/NORW-E9252/2020|EPI_ISL_453585|202 ···················· ···················· ···················· ···················· ···················· ····················
C 6792 hCoV-19/England/NORW-E9261/2020|EPI_ISL_453586|202 ···················· ···················· ···················· ···················· ···················· ····················
C 6793 hCoV-19/England/NORW-E9270/2020|EPI_ISL_453587|202 ···················· ···················· ···················· ···················· ···················· ····················
C 6794 hCoV-19/England/NORW-E92CB/2020|EPI_ISL_453588|202 ···················· ···················· ···················· ···················· ···················· ····················
C 6795 hCoV-19/England/NORW-E9304/2020|EPI_ISL_453591|202 ···················· ···················· ···················· ···················· ···················· ····················
C 6796 hCoV-19/England/NORW-E9331/2020|EPI_ISL_453592|202 ···················· ···················· ···················· ···················· ···················· ····················
C 6797 hCoV-19/England/NORW-E9340/2020|EPI_ISL_453593|202 ···················· ···················· ···················· ···················· ···················· ····················
C 6798 hCoV-19/England/NORW-E935F/2020|EPI_ISL_453594|202 ···················· ···················· ···················· ···················· ···················· ····················
C 6799 hCoV-19/England/NORW-E93AA/2020|EPI_ISL_453595|202 ···················· ···················· ···················· ···················· ···················· ····················
C 6800 hCoV-19/England/NORW-E93B9/2020|EPI_ISL_453596|202 ···················· ···················· ···················· ···················· ···················· ····················
C 6801 hCoV-19/England/NORW-E947A/2020|EPI_ISL_453597|202 ···················· ···················· ···················· ···················· ···················· ····················
C 6802 hCoV-19/England/NORW-E94B6/2020|EPI_ISL_453598|202 ···················· ···················· ···················· ···················· ···················· ····················
C 6803 hCoV-19/England/NORW-E94D4/2020|EPI_ISL_453599|202 ···················· ···················· ···················· ···················· ···················· ····················
C 6804 hCoV-19/USA/NC_200271/2020|EPI_ISL_454810|2020-04 ···················· ···················· ···················· ···················· ···················· ····················
C 6805 hCoV-19/USA/NC_200272/2020|EPI_ISL_454811|2020-04 ···················· ···················· ···················· ···················· ···················· ····················
C 6806 hCoV-19/USA/NC_200278/2020|EPI_ISL_454812|2020-04 ···················· ···················· ···················· ···················· ···················· ····················
C 6807 hCoV-19/USA/NC_200290/2020|EPI_ISL_454813|2020-04 ···················· ···················· ···················· ···················· ···················· ····················
C 6808 hCoV-19/USA/NC_200300/2020|EPI_ISL_454814|2020-03 ···················· ···················· ···················· ···················· ···················· ····················
C 6809 hCoV-19/USA/NC_200306/2020|EPI_ISL_454815|2020-03 ···················· ···················· ···················· ···················· ···················· ····················
C 6810 hCoV-19/USA/NC_200310/2020|EPI_ISL_454816|2020-03 ···················· ···················· ···················· ···················· ···················· ····················
C 6811 hCoV-19/USA/NC_200312/2020|EPI_ISL_454817|2020-03 ···················· ···················· ···················· ···················· ···················· ····················
C 6812 hCoV-19/USA/NC_200318/2020|EPI_ISL_454818|2020-03 ···················· ···················· ···················· ···················· ···················· ····················
C 6813 hCoV-19/USA/NC_200319/2020|EPI_ISL_454819|2020-03 ···················· ···················· ···················· ···················· ···················· ····················
C 6814 hCoV-19/USA/NC_200323/2020|EPI_ISL_454820|2020-03 ···················· ···················· ···················· ···················· ···················· ····················
C 6815 hCoV-19/USA/NC_200325/2020|EPI_ISL_454821|2020-03 ···················· ···················· ···················· ···················· ···················· ····················
C 6816 hCoV-19/USA/NC_200326/2020|EPI_ISL_454822|2020-03 ···················· ···················· ···················· ···················· ···················· ····················
C 6817 hCoV-19/USA/NC_200331/2020|EPI_ISL_454823|2020-03 ···················· ···················· ···················· ···················· ···················· ····················
C 6818 hCoV-19/USA/NC_200330/2020|EPI_ISL_454824|2020-03 ···················· ···················· ···················· ···················· ···················· ····················
C 6819 hCoV-19/USA/NC_200252/2020|EPI_ISL_454825|2020-03 ···················· ···················· ···················· ···················· ···················· ····················
C 6820 hCoV-19/USA/NC_200253/2020|EPI_ISL_454826|2020-03 ···················· ···················· ···················· ···················· ···················· ····················
C 6821 hCoV-19/USA/NC_200340/2020|EPI_ISL_454827|2020-03 ···················· ···················· ···················· ···················· ···················· ····················
C 6822 hCoV-19/USA/NC_200386/2020|EPI_ISL_454828|2020-03 ···················· ···················· ···················· ···················· ···················· ····················
C 6823 hCoV-19/USA/NC_200229/2020|EPI_ISL_454829|2020-04 ···················· ···················· ···················· ···················· ···················· ····················
C 6824 hCoV-19/India/RJ-SMSCOV109/2020|EPI_ISL_454830|202 ···················· ···················· ···················· ···················· ···················· ····················
C 6825 hCoV-19/India/RJ-SMSCOV141/2020|EPI_ISL_454831|202 ···················· ···················· ···················· ···················· ···················· ····················
C 6826 hCoV-19/India/RJ-SMSCOV161/2020|EPI_ISL_454832|202 ···················· ···················· ···················· ···················· ···················· ····················
C 6827 hCoV-19/India/RJ-SMSCOV175/2020|EPI_ISL_454833|202 ···················· ···················· ···················· ···················· ···················· ····················
C 6828 hCoV-19/India/HR-THSTI-BAL-913/2020|EPI_ISL_454865 ···················· ···················· ···················· ···················· ······S··W·········· ····················
C 6829 hCoV-19/India/HR-THSTI-BAL-914/2020|EPI_ISL_454866 ···················· ···················· ···················· ···················· ···················· ····················

| | | | | | | | | | | |
|---|---|---|---|---|---|---|---|---|---|---|
| C | 6830 | hCoV-19/India/HR-THSTI-BAL-918/2020\|EPI_ISL_454867 | ···················· | ···················· | ···················· | ···················· | ···················· | ···················· | | |
| C | 6831 | hCoV-19/Sweden/20-50052/2020\|EPI_ISL_454868\|2020-0 | ···················· | ···················· | ···················· | ···················· | ···················· | ···················· | | |
| C | 6832 | hCoV-19/Sweden/20-50053/2020\|EPI_ISL_454869\|2020-0 | ···················· | ···················· | ···················· | ···················· | ···················· | ···················· | | |
| C | 6833 | hCoV-19/Sweden/20-50091/2020\|EPI_ISL_454874\|2020-0 | ···················· | ···················· | ···················· | ···················· | ···················· | ···················· | | |
| C | 6834 | hCoV-19/Sweden/20-50093/2020\|EPI_ISL_454875\|2020-0 | ···················· | ···················· | ···················· | ···················· | ···················· | ···················· | | |
| C | 6835 | hCoV-19/Sweden/20-50096/2020\|EPI_ISL_454876\|2020-0 | ···················· | ···················· | ···················· | ···················· | ···················· | ···················· | | |
| C | 6836 | hCoV-19/Sweden/20-50098/2020\|EPI_ISL_454877\|2020-0 | ···················· | ···················· | ···················· | ···················· | ···················· | ···················· | | |
| C | 6837 | hCoV-19/Sweden/20-50099/2020\|EPI_ISL_454878\|2020-0 | ···················· | ···················· | ···················· | ···················· | ···················· | ···················· | | |
| C | 6838 | hCoV-19/Sweden/20-50101/2020\|EPI_ISL_454879\|2020-0 | ···················· | ···················· | ···················· | ···················· | ···················· | ···················· | | |
| C | 6839 | hCoV-19/Sweden/20-50104/2020\|EPI_ISL_454880\|2020-0 | ···················· | ···················· | ···················· | ···················· | ···················· | ···················· | | |
| C | 6840 | hCoV-19/Sweden/20-50107/2020\|EPI_ISL_454881\|2020-0 | ···················· | ···················· | ···················· | ···················· | ···················· | ···················· | | |
| C | 6841 | hCoV-19/Sweden/20-50136/2020\|EPI_ISL_454882\|2020-0 | ···················· | ···················· | ···················· | ···················· | ···················· | ···················· | | |
| C | 6842 | hCoV-19/Sweden/20-50139/2020\|EPI_ISL_454883\|2020-0 | ···················· | ···················· | ···················· | ···················· | ···················· | ···················· | | |
| C | 6843 | hCoV-19/Sweden/20-50144/2020\|EPI_ISL_454884\|2020-0 | ··T················· | ···················· | ···················· | ···················· | ···················· | ···················· | | |
| C | 6844 | hCoV-19/Sweden/20-50147/2020\|EPI_ISL_454885\|2020-0 | ··T················· | ···················· | ···················· | ···················· | ···················· | ···················· | | |
| C | 6845 | hCoV-19/Sweden/20-50151/2020\|EPI_ISL_454886\|2020-0 | ···················· | ···················· | ···················· | ···················· | ···················· | ···················· | | |
| C | 6846 | hCoV-19/Sweden/20-50153/2020\|EPI_ISL_454887\|2020-0 | ···················· | ···················· | ···················· | ···················· | ···················· | ···················· | | |
| C | 6847 | hCoV-19/Sweden/20-50155/2020\|EPI_ISL_454888\|2020-0 | ···················· | ···················· | ···················· | ···················· | ···················· | ···················· | | |
| C | 6848 | hCoV-19/Sweden/20-50208/2020\|EPI_ISL_454889\|2020-0 | ···················· | ···················· | ···················· | ···················· | ···················· | ···················· | | |
| C | 6849 | hCoV-19/Sweden/20-50220/2020\|EPI_ISL_454890\|2020-0 | ···················· | ···················· | ···················· | ···················· | ···················· | ···················· | | |
| C | 6850 | hCoV-19/Sweden/20-50222/2020\|EPI_ISL_454891\|2020-0 | ···················· | ···················· | ···················· | ···················· | ···················· | ···················· | | |
| C | 6851 | hCoV-19/Sweden/20-50224/2020\|EPI_ISL_454892\|2020-0 | ···················· | ···················· | ···················· | ···················· | ···················· | ···················· | | |
| C | 6852 | hCoV-19/Sweden/20-50227/2020\|EPI_ISL_454893\|2020-0 | ···················· | ···················· | ···················· | ···················· | ···················· | ···················· | | |
| C | 6853 | hCoV-19/Sweden/20-50248/2020\|EPI_ISL_454894\|2020-0 | ···················· | ···················· | ···················· | ···················· | ···················· | ···················· | | |
| C | 6854 | hCoV-19/Sweden/20-50252/2020\|EPI_ISL_454895\|2020-0 | ···················· | ···················· | ···················· | ···················· | ···················· | ···················· | | |
| C | 6855 | hCoV-19/Sweden/20-50260/2020\|EPI_ISL_454896\|2020-0 | ···················· | ···················· | ···················· | ···················· | ···················· | ···················· | | |
| C | 6856 | hCoV-19/Sweden/20-50283/2020\|EPI_ISL_454897\|2020-0 | ···················· | ···················· | ···················· | ···················· | ···················· | ···················· | | |
| C | 6857 | hCoV-19/Sweden/20-50285/2020\|EPI_ISL_454898\|2020-0 | ···················· | ···················· | ···················· | ···················· | ···················· | ···················· | | |
| C | 6858 | hCoV-19/Sweden/20-50288/2020\|EPI_ISL_454899\|2020-0 | ···················· | ···················· | ···················· | ···················· | ···················· | ···················· | | |
| C | 6859 | hCoV-19/England/EXET-13570C/2020\|EPI_ISL_457090\|20 | ···················· | ···················· | ···················· | ···················· | ···················· | ···················· | | |
| C | 6860 | hCoV-19/England/EXET-13571B/2020\|EPI_ISL_457091\|20 | ···················· | ···················· | ···················· | ···················· | ···················· | ···················· | | |
| C | 6861 | hCoV-19/England/EXET-135739/2020\|EPI_ISL_457092\|20 | ···················· | ···················· | ···················· | ···················· | ···················· | ···················· | | |
| C | 6862 | hCoV-19/England/EXET-135748/2020\|EPI_ISL_457093\|20 | ···················· | ···················· | ···················· | ···················· | ···················· | ···················· | | |
| C | 6863 | hCoV-19/England/EXET-135757/2020\|EPI_ISL_457094\|20 | ···················· | ···················· | ···················· | ···················· | ···················· | ···················· | | |
| C | 6864 | hCoV-19/England/EXET-135766/2020\|EPI_ISL_457095\|20 | ···················· | ···················· | ···················· | ···················· | ···················· | ···················· | | |
| C | 6865 | hCoV-19/England/EXET-135775/2020\|EPI_ISL_457096\|20 | ···················· | ···················· | ···················· | ···················· | ···················· | ···················· | | |
| C | 6866 | hCoV-19/England/EXET-1357B1/2020\|EPI_ISL_457098\|20 | ···················· | ···················· | ···················· | ···················· | ···················· | ···················· | | |
| C | 6867 | hCoV-19/England/EXET-1357C0/2020\|EPI_ISL_457099\|20 | ···················· | ···················· | ···················· | ···················· | ···················· | ···················· | | |
| C | 6868 | hCoV-19/Belgium/Rega-0329230/2020\|EPI_ISL_458200\|2 | ···················· | ···················· | ···················· | ···················· | ···················· | ···················· | | |
| C | 6869 | hCoV-19/Belgium/Rega-0329231/2020\|EPI_ISL_458201\|2 | ···················· | ···················· | ···················· | ···················· | ···················· | ···················· | | |
| C | 6870 | hCoV-19/Belgium/Rega-0329232/2020\|EPI_ISL_458202\|2 | ···················· | ···················· | ···················· | ···················· | ···················· | ···················· | | |
| C | 6871 | hCoV-19/Belgium/Rega-0329233/2020\|EPI_ISL_458203\|2 | ···················· | ···················· | ···················· | ···················· | ···················· | ···················· | | |
| C | 6872 | hCoV-19/Belgium/Rega-0329234/2020\|EPI_ISL_458204\|2 | ···················· | ···················· | ···················· | ···················· | ···················· | ···················· | | |
| C | 6873 | hCoV-19/Belgium/Rega-0329235/2020\|EPI_ISL_458205\|2 | ···················· | ···················· | ···················· | ···················· | ···················· | ···················· | | |
| C | 6874 | hCoV-19/Belgium/Rega-0329236/2020\|EPI_ISL_458206\|2 | ···················· | ···················· | ···················· | ···················· | ···················· | ···················· | | |
| C | 6875 | hCoV-19/Scotland/GCVR-170486/2020\|EPI_ISL_459506\|2 | ···················· | ···················· | ···················· | ···················· | ···················· | ···················· | | |
| C | 6876 | hCoV-19/Scotland/GCVR-170574/2020\|EPI_ISL_459508\|2 | ···················· | ···················· | ···················· | ···················· | ···················· | ···················· | | |
| C | 6877 | hCoV-19/Scotland/GCVR-1710FA/2020\|EPI_ISL_459509\|2 | ···················· | ···················· | ···················· | ···················· | ···················· | ···················· | | |
| C | 6878 | hCoV-19/Scotland/GCVR-171115/2020\|EPI_ISL_459510\|2 | ···················· | ···················· | ···················· | ···················· | ···················· | ···················· | | |
| C | 6879 | hCoV-19/Scotland/GCVR-170310/2020\|EPI_ISL_459513\|2 | ···················· | ···················· | ···················· | ···················· | ···················· | ···················· | | |
| C | 6880 | hCoV-19/Scotland/GCVR-170AFD/2020\|EPI_ISL_459514\|2 | ···················· | ···················· | ···················· | ···················· | ···················· | ···················· | | |
| C | 6881 | hCoV-19/Scotland/GCVR-1711F7/2020\|EPI_ISL_459517\|2 | ···················· | ···················· | ···················· | ···················· | ···················· | ···················· | | |
| C | 6882 | hCoV-19/Scotland/GCVR-170680/2020\|EPI_ISL_459518\|2 | ···················· | ···················· | ···················· | ···················· | ···················· | ···················· | | |
| C | 6883 | hCoV-19/Scotland/GCVR-1705A1/2020\|EPI_ISL_459519\|2 | ···················· | ···················· | ···················· | ···················· | ···················· | ···················· | | |
| C | 6884 | hCoV-19/Scotland/GCVR-1710BE/2020\|EPI_ISL_459520\|2 | ···················· | ···················· | ···················· | ···················· | ···················· | ···················· | | |
| C | 6885 | hCoV-19/Scotland/GCVR-170F76/2020\|EPI_ISL_459521\|2 | ···················· | ···················A | ···················· | ···················· | ···················· | ···················· | 1 | |
| C | 6886 | hCoV-19/Scotland/GCVR-1711CA/2020\|EPI_ISL_459522\|2 | ···················· | ···················· | ···················· | ···················· | ···················· | ···················· | | |
| C | 6887 | hCoV-19/Scotland/GCVR-170213/2020\|EPI_ISL_459524\|2 | ···················· | ···················· | ···················· | ···················· | ···················· | ···················· | | |
| C | 6888 | hCoV-19/Scotland/GCVR-16FF10/2020\|EPI_ISL_459525\|2 | ···················· | ···················· | ···················· | ···················· | ···················· | ···················· | | |
| C | 6889 | hCoV-19/Scotland/GCVR-17044A/2020\|EPI_ISL_459526\|2 | ···················· | ···················· | ···················· | ···················· | ···················· | ···················· | | |
| C | 6890 | hCoV-19/Scotland/GCVR-170C06/2020\|EPI_ISL_459527\|2 | ···················· | ·A·················· | ···················· | ···················· | ···················· | ···················· | | |
| C | 6891 | hCoV-19/Scotland/GCVR-1712F4/2020\|EPI_ISL_459528\|2 | ···················· | ···················· | ···················· | ···················· | ···················· | ···················· | | |
| C | 6892 | hCoV-19/Scotland/GCVR-170B27/2020\|EPI_ISL_459529\|2 | ···················· | ···················· | ···················· | ···················· | ···················· | ···················· | | |
| C | 6893 | hCoV-19/Scotland/GCVR-171203/2020\|EPI_ISL_459530\|2 | ···················· | ···················· | ···················· | ···················· | ···················· | ···················· | | |
| C | 6894 | hCoV-19/Scotland/GCVR-1700BF/2020\|EPI_ISL_459532\|2 | ···················· | ·A·················· | ···················· | ···················· | ···················· | ···················· | | |
| C | 6895 | hCoV-19/Scotland/GCVR-170091/2020\|EPI_ISL_459533\|2 | ···················· | ···················· | ···················· | ···················· | ···················· | ···················· | | |
| C | 6896 | hCoV-19/Scotland/GCVR-170996/2020\|EPI_ISL_459534\|2 | ···················· | ···················· | ···················· | ···················· | ···················· | ···················· | | |
| C | 6897 | hCoV-19/Scotland/GCVR-1713A6/2020\|EPI_ISL_459535\|2 | ···················· | ···················· | ···················· | ···················· | ···················· | ···················· | | |
| C | 6898 | hCoV-19/Scotland/GCVR-17018F/2020\|EPI_ISL_459536\|2 | ···················· | ···················· | ···················· | ···················· | ···················· | ···················· | | |
| C | 6899 | hCoV-19/Scotland/GCVR-1705CF/2020\|EPI_ISL_459700\|2 | ···················· | ···················· | ···················· | ···················· | ···················· | ···················· | | |
| C | 6900 | hCoV-19/Scotland/GCVR-171036/2020\|EPI_ISL_459701\|2 | ···················· | ·A·················· | ···················· | ···················· | ···················· | ···················· | | |
| C | 6901 | hCoV-19/Scotland/GCVR-16FE13/2020\|EPI_ISL_459702\|2 | ···················· | ···················· | ···················· | ···················· | ···················· | ···················· | | |
| C | 6902 | hCoV-19/England/NORW-E90FE/2020\|EPI_ISL_461820\|202 | ···················· | ···················· | ···················· | ···················· | ···················· | ···················· | | |
| C | 6904 | hCoV-19/England/NORW-E9119/2020\|EPI_ISL_461822\|202 | ···················· | ···················· | ···················· | ···················· | NNN················· | ···················· | | 1 |
| C | 6905 | hCoV-19/England/NORW-E9137/2020\|EPI_ISL_461823\|202 | ···················· | ···················· | ···················· | ···················· | ···················· | ···················· | | |
| C | 6906 | hCoV-19/England/NORW-E9182/2020\|EPI_ISL_461825\|202 | ···················· | ···················· | ···················· | ···················· | NNN················· | ···················· | | 1 |
| C | 6907 | hCoV-19/England/NORW-E9207/2020\|EPI_ISL_461827\|202 | ···················· | ···················· | ···················· | ···················· | ···················· | ···················· | | |
| C | 6908 | hCoV-19/England/NORW-E9322/2020\|EPI_ISL_461828\|202 | ···················· | ···················· | ···················· | ···················· | ···················· | ···················· | | |

| | | | | | | | | |
|---|---|---|---|---|---|---|---|---|
| C | 6911 hCoV-19/England/NORW-E9999/2020\|EPI_ISL_461839\|202 | ···················· | ···················· | ···················· | ···················· | ···················· | ···················· | |
| C | 6913 hCoV-19/Northern Ireland/NIRE-FAFDF/2020\|EPI_ISL_4 | ···················· | ···················· | ···················· | ···················· | ···················· | ···················· | |
| C | 6927 hCoV-19/Northern Ireland/NIRE-FB721/2020\|EPI_ISL_4 | ···················· | ···················· | ···················· | ···················· | ···················· | ···················· | |
| C | 6928 hCoV-19/Northern Ireland/NIRE-FB730/2020\|EPI_ISL_4 | ···················· | ···················· | ···················· | ···················· | ···················· | ···················· | |
| C | 6933 hCoV-19/India/MH-NIV-12024/2020\|EPI_ISL_452200\|202 | ···················· | ···················· | ···················· | ·········------····· | ···················· | ···················· | 1 |
| C | 6934 hCoV-19/India/MH-NIV-7511/2020\|EPI_ISL_452208\|2020 | ···················· | ···················· | ···················· | ···················· | ···················· | ···················· | |
| C | 6935 hCoV-19/India/MH-NIV-7830/2020\|EPI_ISL_452210\|2020 | ···················· | ···················· | ···················· | ···················· | ···················· | ···················· | |
| C | 6936 hCoV-19/India/MH-NIV-9157/2020\|EPI_ISL_452211\|2020 | ···················· | ···················· | ···················· | ···················· | ···················· | ···················· | |
| C | 6937 hCoV-19/England/NORT-291855/2020\|EPI_ISL_453542\|20 | ···················· | ···················· | ···················· | ···················· | ···················· | ···················· | |
| C | 6938 hCoV-19/USA/NC_200242/2020\|EPI_ISL_454800\|2020-03 | ···················· | ···················· | ···················· | ···················· | ···················· | ···················· | |
| C | 6939 hCoV-19/USA/NC_200243/2020\|EPI_ISL_454801\|2020-03 | ···················· | ···················· | ···················· | ···················· | ···················· | ···················· | |
| C | 6940 hCoV-19/USA/NC_200244/2020\|EPI_ISL_454802\|2020-03 | ···················· | ···················· | ···················· | ···················· | ···················· | ···················· | |
| C | 6941 hCoV-19/USA/NC_200245/2020\|EPI_ISL_454803\|2020-03 | ···················· | ···················· | ···················· | ···················· | ···················· | ···················· | |
| C | 6942 hCoV-19/USA/NC_200247/2020\|EPI_ISL_454804\|2020-03 | ···················· | ···················· | ···················· | ···················· | ···················· | ···················· | |
| C | 6943 hCoV-19/USA/NC_200254/2020\|EPI_ISL_454805\|2020-03 | ···················· | ···················· | ···················· | ···················· | ···················· | ···················· | |
| C | 6944 hCoV-19/USA/NC_200255/2020\|EPI_ISL_454806\|2020-03 | ···················· | ···················· | ···················· | ···················· | ···················· | ···················· | |
| C | 6945 hCoV-19/USA/NC_200256/2020\|EPI_ISL_454807\|2020-03 | ···················· | ···················· | ···················· | ···················· | ···················· | ···················· | |
| C | 6946 hCoV-19/USA/NC_200259/2020\|EPI_ISL_454808\|2020-03 | ···················· | ···················· | ···················· | ···················· | ···················· | ···················· | |
| C | 6947 hCoV-19/USA/NC_200264/2020\|EPI_ISL_454809\|2020-04 | ···················· | ···················· | ···················· | ···················· | ···················· | ···················· | |
| C | 6948 hCoV-19/India/HR-THSTI-BAL-39/2020\|EPI_ISL_454858\| | ···················· | ···················· | ···················· | ···················· | ···················· | ···················· | |
| C | 6949 hCoV-19/India/HR-THSTI-BAL-231/2020\|EPI_ISL_454862 | ···················· | ···················· | ···················· | ···················· | ···················· | ···················· | |
| C | 6950 hCoV-19/Sweden/20-50055/2020\|EPI_ISL_454870\|2020-0 | ···················· | ···················· | ···················· | ···················· | ···················· | ···················· | |
| C | 6951 hCoV-19/Sweden/20-50088/2020\|EPI_ISL_454871\|2020-0 | ···················· | ···················· | ···················· | ···················· | ···················· | ···················· | |
| C | 6952 hCoV-19/Sweden/20-50089/2020\|EPI_ISL_454872\|2020-0 | ···················· | ···················· | ···················· | ···················· | ···················· | ···················· | |
| C | 6953 hCoV-19/Sweden/20-50090/2020\|EPI_ISL_454873\|2020-0 | ···················· | ···················· | ···················· | ···················· | ···················· | ···················· | |
| C | 6954 hCoV-19/Scotland/EDB5368/2020\|EPI_ISL_457009\|2020- | ···················· | ···················· | ···················· | ···················· | ···················· | ···················· | |
| C | 6955 hCoV-19/Scotland/EDB5370/2020\|EPI_ISL_457010\|2020- | ···················· | ···················· | ···················· | ···················· | ···················· | ···················· | |
| C | 6956 hCoV-19/Scotland/EDB5373/2020\|EPI_ISL_457011\|2020- | ···················· | ···················· | ···················· | ···················· | ···················· | ···················· | |
| C | 6958 hCoV-19/Scotland/EDB554/2020\|EPI_ISL_457013\|2020-0 | ···················· | ···················· | ···················· | ···················· | ···················· | ···················· | |
| C | 6959 hCoV-19/Scotland/EDB704/2020\|EPI_ISL_457015\|2020-0 | ···················· | ···················· | ···················· | ···················· | ···················· | ···················· | |
| C | 6960 hCoV-19/Scotland/EDB805/2020\|EPI_ISL_457018\|2020-0 | ···················· | ···················· | ···················· | ···················· | ···················· | ···················· | |
| C | 6961 hCoV-19/Scotland/EDB808/2020\|EPI_ISL_457019\|2020-0 | ···················· | ···················· | ···················· | ···················· | ···················· | ···················· | |
| C | 6962 hCoV-19/Scotland/EDB816/2020\|EPI_ISL_457020\|2020-0 | ···················· | ···················· | ···················· | ···················· | ···················· | ···················· | |
| C | 6963 hCoV-19/Scotland/EDB890/2020\|EPI_ISL_457021\|2020-0 | ···················· | ···················· | ···················· | ···················· | ···················· | ···················· | |
| C | 6964 hCoV-19/Scotland/EDB949/2020\|EPI_ISL_457023\|2020-0 | ···················· | ···················· | ···················· | ···················· | ···················· | ···················· | |
| C | 6965 hCoV-19/England/EXET-134FAB/2020\|EPI_ISL_457025\|20 | ···················· | ···················· | ···················· | ···················· | ···················· | ···················· | |
| C | 6966 hCoV-19/England/EXET-134FC9/2020\|EPI_ISL_457026\|20 | ···················· | ···················· | ···················· | ···················· | ···················· | ···················· | |
| C | 6967 hCoV-19/England/EXET-134FD8/2020\|EPI_ISL_457027\|20 | ···················· | ···················· | ···················· | ···················· | ···················· | ···················· | |
| C | 6968 hCoV-19/England/EXET-135098/2020\|EPI_ISL_457028\|20 | ···················· | ···················· | ···················· | ···················· | ···················· | ···················· | |
| C | 6969 hCoV-19/England/EXET-1350A7/2020\|EPI_ISL_457029\|20 | ···················· | ···················· | ···················· | ···················· | ···················· | ···················· | |
| C | 6970 hCoV-19/England/EXET-1350D4/2020\|EPI_ISL_457030\|20 | ···················· | ···················· | ···················· | ···················· | ···················· | ···················· | |
| C | 6971 hCoV-19/England/EXET-1350E3/2020\|EPI_ISL_457031\|20 | ···················· | ···················· | ···················· | ···················· | ···················· | ···················· | |
| C | 6972 hCoV-19/England/EXET-1351FF/2020\|EPI_ISL_457035\|20 | ···················· | ···················· | ···················· | ···················· | ···················· | ···················· | |
| C | 6973 hCoV-19/England/EXET-13520B/2020\|EPI_ISL_457036\|20 | ···················· | ···················· | ···················· | ···················· | ···················· | ···················· | |
| C | 6974 hCoV-19/England/EXET-135238/2020\|EPI_ISL_457038\|20 | ···················· | ···················· | ···················· | ···················· | ···················· | ···················· | |
| C | 6975 hCoV-19/England/EXET-135265/2020\|EPI_ISL_457040\|20 | ···················· | ···················· | ···················· | ···················· | ···················· | ···················· | |
| C | 6976 hCoV-19/England/EXET-135274/2020\|EPI_ISL_457041\|20 | ···················· | ···················· | ···················· | ···················· | ···················· | ···················· | |
| C | 6977 hCoV-19/England/EXET-1352A1/2020\|EPI_ISL_457043\|20 | ···················· | ···················· | ···················· | ···················· | ···················· | ···················· | |
| C | 6978 hCoV-19/England/EXET-1352B0/2020\|EPI_ISL_457044\|20 | ···················· | ···················· | ···················· | ···················· | ···················· | ···················· | |
| C | 6979 hCoV-19/England/EXET-1352FC/2020\|EPI_ISL_457047\|20 | ···················· | ···················· | ···················· | ···················· | ···················· | ···················· | |
| C | 6980 hCoV-19/England/EXET-135335/2020\|EPI_ISL_457049\|20 | ···················· | ···················· | ···················· | ···················· | ···················· | ···················· | |
| C | 6981 hCoV-19/England/EXET-135371/2020\|EPI_ISL_457052\|20 | ···················· | ···················· | ···················· | ···················· | ···················· | ···················· | |
| C | 6982 hCoV-19/England/EXET-1353CC/2020\|EPI_ISL_457053\|20 | ···················· | ···················· | ···················· | ···················· | ···················· | ···················· | |
| C | 6983 hCoV-19/England/EXET-1353DB/2020\|EPI_ISL_457054\|20 | ···················· | ···················· | ···················· | ···················· | ···················· | ···················· | |
| C | 6984 hCoV-19/England/EXET-1353EA/2020\|EPI_ISL_457055\|20 | ···················· | ···················· | ···················· | ···················· | ···················· | ···················· | |
| C | 6985 hCoV-19/England/EXET-1353F9/2020\|EPI_ISL_457056\|20 | ···················· | ···················· | ···················· | ···················· | ···················· | ···················· | |
| C | 6986 hCoV-19/England/EXET-135405/2020\|EPI_ISL_457057\|20 | ···················· | ···················· | ···················· | ···················· | ···················· | ···················· | |
| C | 6987 hCoV-19/England/EXET-135423/2020\|EPI_ISL_457058\|20 | ···················· | ···················· | ···················· | ···················· | ···················· | ···················· | |
| C | 6988 hCoV-19/England/EXET-135432/2020\|EPI_ISL_457059\|20 | ···················· | ···················· | ···················· | ···················· | ···················· | ···················· | |
| C | 6989 hCoV-19/England/EXET-135441/2020\|EPI_ISL_457060\|20 | ···················· | ···················· | ···················· | ···················· | ···················· | ···················· | |
| C | 6990 hCoV-19/England/EXET-135450/2020\|EPI_ISL_457061\|20 | ···················· | ···················· | ···················· | ···················· | ···················· | ···················· | |
| C | 6991 hCoV-19/England/EXET-13546F/2020\|EPI_ISL_457062\|20 | ···················· | ···················· | ···················· | ···················· | ···················· | ···················· | |
| C | 6992 hCoV-19/England/EXET-13547E/2020\|EPI_ISL_457063\|20 | ···················· | ···················· | ···················· | ···················· | ···················· | ···················· | |
| C | 6993 hCoV-19/England/EXET-13548D/2020\|EPI_ISL_457064\|20 | ···················· | ···················· | ···················· | ···················· | ···················· | ···················· | |
| C | 6994 hCoV-19/England/EXET-13549C/2020\|EPI_ISL_457065\|20 | ···················· | ···················· | ···················· | ···················· | ···················· | ···················· | |
| C | 6995 hCoV-19/England/EXET-1354AB/2020\|EPI_ISL_457066\|20 | ···················· | ···················· | ···················· | ···················· | ···················· | ···················· | |
| C | 6996 hCoV-19/England/EXET-1354C9/2020\|EPI_ISL_457067\|20 | ···················· | ···················· | ···················· | ···················· | ···················· | ···················· | |
| C | 6997 hCoV-19/England/EXET-135502/2020\|EPI_ISL_457068\|20 | ···················· | ···················· | ···················· | ···················· | ···················· | ···················· | |
| C | 6998 hCoV-19/England/EXET-135511/2020\|EPI_ISL_457069\|20 | ···················· | ···················· | ···················· | ···················· | ···················· | ···················· | |
| C | 6999 hCoV-19/England/EXET-13555D/2020\|EPI_ISL_457070\|20 | ···················· | ···················· | ···················· | ···················· | ···················· | ···················· | |
| C | 7001 hCoV-19/England/EXET-1355B7/2020\|EPI_ISL_457073\|20 | ···················· | ···················· | ···················· | ···················· | ···················· | ···················· | |
| C | 7002 hCoV-19/England/EXET-1355C6/2020\|EPI_ISL_457074\|20 | ···················· | ···················· | ···················· | ···················· | ···················· | ···················· | |
| C | 7003 hCoV-19/England/EXET-1355D5/2020\|EPI_ISL_457075\|20 | ···················· | ···················· | ···················· | ···················· | ···················· | ···················· | |
| C | 7004 hCoV-19/England/EXET-13560F/2020\|EPI_ISL_457076\|20 | ···················· | ···················· | ···················· | ···················· | ···················· | ···················· | |
| C | 7005 hCoV-19/England/EXET-13561E/2020\|EPI_ISL_457077\|20 | ···················· | ···················· | ···················· | ···················· | ···················· | ···················· | |
| C | 7006 hCoV-19/England/EXET-13562D/2020\|EPI_ISL_457078\|20 | ···················· | ···················· | ···················· | ···················· | ···················· | ···················· | |
| C | 7007 hCoV-19/England/EXET-13563C/2020\|EPI_ISL_457079\|20 | ···················· | ···················· | ···················· | ···················· | ···················· | ···················· | |
| C | 7008 hCoV-19/England/EXET-13565A/2020\|EPI_ISL_457081\|20 | ···················· | ·············A······ | ···················· | ···················· | ···················· | ···················· | |

| | | | | | | | | |
|---|---|---|---|---|---|---|---|---|
| C | 7009 | hCoV-19/England/EXET-135687/2020\|EPI_ISL_457083\|20 | ···················· | ···················· | ···················· | ···················· | ···················· | ···················· |
| C | 7010 | hCoV-19/England/EXET-1356B4/2020\|EPI_ISL_457086\|20 | ···················· | ···················· | ···················· | ···················· | ···················· | ···················· |
| C | 7011 | hCoV-19/England/EXET-1356D2/2020\|EPI_ISL_457087\|20 | ···················· | ···················· | ···················· | ···················· | ···················· | ···················· |
| C | 7012 | hCoV-19/England/EXET-1356E1/2020\|EPI_ISL_457088\|20 | ···················· | ···················· | ···················· | ···················· | ···················· | ···················· |
| C | 7013 | hCoV-19/England/EXET-1356F0/2020\|EPI_ISL_457089\|20 | ···················· | ···················· | ···················· | ···················· | ···················· | ···················· |
| C | 7014 | hCoV-19/England/BRIS-12AA3E/2020\|EPI_ISL_458339\|20 | ···················· | ···················· | ···················· | ···················· | ···················· | ···················· |
| C | 7015 | hCoV-19/England/BRIS-129E9D/2020\|EPI_ISL_458341\|20 | ···················· | ···················· | ···················· | ···················· | ···················· | ···················· |
| C | 7016 | hCoV-19/England/BRIS-12A2FA/2020\|EPI_ISL_458343\|20 | ···················· | ·············A······ | ···················· | ···················· | ···················· | ···················· |
| C | 7017 | hCoV-19/England/BRIS-12ADEA/2020\|EPI_ISL_458344\|20 | ···················· | ···················· | ···················· | ···················· | ···················· | ···················· |
| C | 7018 | hCoV-19/England/BRIS-12A61C/2020\|EPI_ISL_458345\|20 | ···················· | ···················· | ···················· | ···················· | ···················· | ···················· |
| C | 7019 | hCoV-19/England/BRIS-129F21/2020\|EPI_ISL_458348\|20 | ···················· | ···················· | ···················· | ···················· | ···················· | ···················· |
| C | 7020 | hCoV-19/England/BRIS-12A184/2020\|EPI_ISL_458349\|20 | ···················· | ···················· | ···················· | ···················· | ···················· | ···················· |
| C | 7021 | hCoV-19/England/BRIS-12A9C8/2020\|EPI_ISL_458350\|20 | ···················· | ···················· | ···················· | ···················· | ···················· | ···················· |
| C | 7022 | hCoV-19/England/BRIS-12AC56/2020\|EPI_ISL_458351\|20 | ···················· | ···················· | ···················· | ···················· | ···················· | ···················· |
| C | 7023 | hCoV-19/England/BRIS-12AB2C/2020\|EPI_ISL_458352\|20 | ···················· | ···················· | ···················· | ···················· | ···················· | ···················· |
| C | 7024 | hCoV-19/England/BRIS-12A0A5/2020\|EPI_ISL_458353\|20 | ···················· | ···················· | ···················· | ···················· | ···················· | ···················· |
| C | 7025 | hCoV-19/England/BRIS-12AEBA/2020\|EPI_ISL_458354\|20 | ···················· | ···················· | ···················· | ···················· | ···················· | ···················· |
| C | 7026 | hCoV-19/England/BRIS-12AE05/2020\|EPI_ISL_458355\|20 | ···················· | ···················· | ···················· | ···················· | ···················· | ···················· |
| C | 7027 | hCoV-19/England/BRIS-1298DB/2020\|EPI_ISL_458357\|20 | ···················· | ···················· | ···················· | ···················· | ···················· | ···················· |
| C | 7028 | hCoV-19/England/BRIS-12A421/2020\|EPI_ISL_458358\|20 | ···················· | ···················· | ···················· | ···················· | ···················· | ···················· |
| C | 7029 | hCoV-19/England/BRIS-129D27/2020\|EPI_ISL_458359\|20 | ···················· | ···················· | ···················· | ···················· | ···················· | ···················· |
| C | 7030 | hCoV-19/England/BRIS-12AD71/2020\|EPI_ISL_458360\|20 | ···················· | ···················· | ···················· | ···················· | ···················· | ···················· |
| C | 7031 | hCoV-19/England/BRIS-129C2A/2020\|EPI_ISL_458361\|20 | ···················· | ···················· | ···················· | ···················· | ···················· | ···················· |
| C | 7032 | hCoV-19/England/BRIS-12A403/2020\|EPI_ISL_458362\|20 | ···················· | ···················· | ···················· | ···················· | ···················· | ···················· |
| C | 7033 | hCoV-19/England/BRIS-12AAB6/2020\|EPI_ISL_458363\|20 | ···················· | ···················· | ···················· | ···················· | ···················· | ···················· |
| C | 7034 | hCoV-19/England/BRIS-129871/2020\|EPI_ISL_458365\|20 | ···················· | ···················· | ···················· | ···················· | ···················· | ···················· |
| C | 7035 | hCoV-19/England/BRIS-12ACFC/2020\|EPI_ISL_458367\|20 | ···················· | ···················· | ···················· | ···················· | ···················· | ···················· |
| C | 7036 | hCoV-19/England/BRIS-129CFD/2020\|EPI_ISL_458368\|20 | ···················· | ···················· | ···················· | ···················· | ···················· | ···················· |
| C | 7037 | hCoV-19/England/BRIS-129B5A/2020\|EPI_ISL_458370\|20 | ···················· | ···A················ | ···················· | ···················· | ···················· | ···················· |
| C | 7038 | hCoV-19/England/BRIS-129E42/2020\|EPI_ISL_458371\|20 | ···················· | ···················· | ···················· | ···················· | ···················· | ···················· |
| C | 7039 | hCoV-19/England/BRIS-129932/2020\|EPI_ISL_458372\|20 | ···················· | ···················· | ···················· | ···················· | ···················· | ···················· |
| C | 7040 | hCoV-19/England/BRIS-129D81/2020\|EPI_ISL_458375\|20 | ···················· | ···················· | ···················· | ···················· | ···················· | ···················· |
| C | 7041 | hCoV-19/England/BRIS-129EAC/2020\|EPI_ISL_458377\|20 | ···················· | ···················· | ···················· | ···················· | ···················· | ···················· |
| C | 7042 | hCoV-19/England/BRIS-12A913/2020\|EPI_ISL_458379\|20 | ···················· | ···················· | ···················· | ···················· | ···················· | ···················· |
| C | 7043 | hCoV-19/England/BRIS-12A1A2/2020\|EPI_ISL_458381\|20 | ···················· | ···················· | ···················· | ···················· | ···················· | ···················· |
| C | 7044 | hCoV-19/England/BRIS-12A4C7/2020\|EPI_ISL_458382\|20 | ···················· | ···················· | ···················· | ···················· | ···················· | ···················· |
| C | 7045 | hCoV-19/England/BRIS-129FE5/2020\|EPI_ISL_458384\|20 | ···················· | ···················· | ···················· | ···················· | ···················· | ···················· |
| C | 7046 | hCoV-19/England/BRIS-12A54C/2020\|EPI_ISL_458385\|20 | ···················· | ···················· | ···················· | ···················· | ···················· | ···················· |
| C | 7047 | hCoV-19/England/BRIS-129B96/2020\|EPI_ISL_458391\|20 | ···················· | ···················· | ···················· | ···················· | ···················· | ···················· |
| C | 7048 | hCoV-19/England/BRIS-12A9E6/2020\|EPI_ISL_458394\|20 | ···················· | ···················· | ···················· | ···················· | ···················· | ···················· |
| C | 7049 | hCoV-19/England/BRIS-12A236/2020\|EPI_ISL_458395\|20 | ···················· | ·············A······ | ···················· | ···················· | ···················· | ···················· |
| C | 7050 | hCoV-19/England/BRIS-12A2DC/2020\|EPI_ISL_458396\|20 | ···················· | ···················· | ···················· | ···················· | ···················· | ···················· |
| C | 7051 | hCoV-19/England/BRIS-12AF8A/2020\|EPI_ISL_458397\|20 | ···················· | ···················· | ···················· | ···················· | ···················· | ···················· |
| C | 7052 | hCoV-19/Scotland/GCVR-171221/2020\|EPI_ISL_459600\|2 | ···················· | ···················· | ···················· | ···················· | ···················· | ···················· |
| C | 7053 | hCoV-19/Scotland/GCVR-170B45/2020\|EPI_ISL_459602\|2 | ···················· | ···················· | ···················· | ···················· | ···················· | ···················· |
| C | 7054 | hCoV-19/Scotland/GCVR-1708D5/2020\|EPI_ISL_459603\|2 | ···················· | ···················· | ···················· | ···················· | ···················· | ···················· |
| C | 7055 | hCoV-19/Scotland/GCVR-170BFA/2020\|EPI_ISL_459605\|2 | ···················· | ·A·················· | ···················· | ···················· | ···················· | ···················· |
| C | 7056 | hCoV-19/Scotland/GCVR-170C15/2020\|EPI_ISL_459606\|2 | ···················· | ···················· | ···················· | ···················· | ···················· | ···················· |
| C | 7057 | hCoV-19/Scotland/GCVR-170B18/2020\|EPI_ISL_459607\|2 | ···················· | ···················· | ···················· | ···················· | ···················· | ···················· |
| C | 7058 | hCoV-19/Scotland/GCVR-170C60/2020\|EPI_ISL_459608\|2 | ···················· | ···················· | ···················· | ···················· | ···················· | ···················· |
| C | 7059 | hCoV-19/Scotland/GCVR-170D4F/2020\|EPI_ISL_459609\|2 | ···················· | ···················· | ···················· | ···················· | ···················· | ···················· |
| C | 7060 | hCoV-19/Scotland/GCVR-170BEB/2020\|EPI_ISL_459610\|2 | ···················· | ·A·················· | ···················· | ···················· | ···················· | ···················· |
| C | 7061 | hCoV-19/Scotland/GCVR-1701E9/2020\|EPI_ISL_459613\|2 | ···················· | ···················· | ···················· | ···················· | ···················· | ···················· |
| C | 7062 | hCoV-19/Scotland/GCVR-170617/2020\|EPI_ISL_459614\|2 | ···················· | ···················· | ···················· | ···················· | ···················· | ···················· |
| C | 7063 | hCoV-19/Scotland/GCVR-171124/2020\|EPI_ISL_459616\|2 | ···················· | ···················· | ···················· | ···················· | ···················· | ···················· |
| C | 7064 | hCoV-19/Scotland/GCVR-170152/2020\|EPI_ISL_459617\|2 | ···················· | ·············A······ | ···················· | ···················· | ···················· | ···················· |
| C | 7065 | hCoV-19/Scotland/GCVR-170F94/2020\|EPI_ISL_459619\|2 | ···················· | ···················· | ···················· | ···················· | ···················· | ···················· |
| C | 7066 | hCoV-19/Scotland/GCVR-1712B8/2020\|EPI_ISL_459621\|2 | ···················· | ···················· | ···················· | ···················· | ···················· | ···················· |
| C | 7067 | hCoV-19/Scotland/GCVR-17036B/2020\|EPI_ISL_459623\|2 | ···················· | ···················· | ···················· | ···················· | ···················· | ···················· |
| C | 7068 | hCoV-19/Scotland/GCVR-1708F3/2020\|EPI_ISL_459624\|2 | ···················· | ···················· | ···················· | ···················· | ···················· | ···················· |
| C | 7069 | hCoV-19/Scotland/GCVR-17000A/2020\|EPI_ISL_459625\|2 | ···················· | ···················· | ···················· | ···················· | ···················· | ···················· |
| C | 7070 | hCoV-19/Scotland/GCVR-171072/2020\|EPI_ISL_459670\|2 | ···················· | ···················· | ···················· | ···················· | ···················· | ···················· |
| C | 7071 | hCoV-19/Scotland/GCVR-17133D/2020\|EPI_ISL_459672\|2 | ···················· | ···················· | ···················· | ···················· | ···················· | ···················· |
| C | 7072 | hCoV-19/Scotland/GCVR-17129A/2020\|EPI_ISL_459678\|2 | ···················· | ···················· | ···················· | ···················· | ···················· | ···················· |
| C | 7073 | hCoV-19/Scotland/GCVR-17040E/2020\|EPI_ISL_459679\|2 | ···················· | ···················· | ···················· | ···················· | ···················· | ···················· |
| C | 7074 | hCoV-19/Scotland/GCVR-1712C7/2020\|EPI_ISL_459680\|2 | ···················· | ···················· | ···················· | ···················· | ···················· | ···················· |
| C | 7075 | hCoV-19/Scotland/GCVR-1709C3/2020\|EPI_ISL_459681\|2 | ···················· | ·A·················· | ···················· | ···················· | ···················· | ···················· |
| C | 7076 | hCoV-19/Scotland/GCVR-171090/2020\|EPI_ISL_459682\|2 | ···················· | ···················· | ···················· | ···················· | ···················· | ···················· |
| C | 7077 | hCoV-19/Scotland/GCVR-1713F1/2020\|EPI_ISL_459684\|2 | ···················· | ···················· | ···················· | ···················· | ···················· | ···················· |
| C | 7078 | hCoV-19/Scotland/GCVR-17027D/2020\|EPI_ISL_459686\|2 | ···················· | ···················· | ···················· | ···················· | ···················· | ···················· |
| C | 7079 | hCoV-19/Scotland/GCVR-1705ED/2020\|EPI_ISL_459688\|2 | ···················· | ···················· | ···················· | ···················· | ···················· | ···················· |
| C | 7080 | hCoV-19/Scotland/GCVR-1710AF/2020\|EPI_ISL_459689\|2 | ···················· | ···················· | ···················· | ···················· | ···················· | ···················· |
| C | 7081 | hCoV-19/Scotland/GCVR-170301/2020\|EPI_ISL_459690\|2 | ···················· | ···················· | ···················· | ···················· | ···················· | ···················· |
| C | 7082 | hCoV-19/Scotland/GCVR-170037/2020\|EPI_ISL_459691\|2 | ···················· | ·A·················· | ···················· | ···················· | ···················· | ···················· |
| C | 7083 | hCoV-19/Scotland/GCVR-17126D/2020\|EPI_ISL_459692\|2 | ···················· | ···················· | ···················· | ···················· | ···················· | ···················· |
| C | 7084 | hCoV-19/Scotland/GCVR-171009/2020\|EPI_ISL_459693\|2 | ···················· | ···················· | ···················· | ···················· | ···················· | ···················· |
| C | 7085 | hCoV-19/Scotland/GCVR-170969/2020\|EPI_ISL_459694\|2 | ···················· | ·A·················· | ···················· | ···················· | ···················· | ···················· |
| C | 7086 | hCoV-19/Scotland/GCVR-171027/2020\|EPI_ISL_459695\|2 | ···················· | ·A·················· | ···················· | ···················· | ···················· | ···················· |

| | | | | | | | |
|---|---|---|---|---|---|---|---|
| C | 7087 hCoV-19/Scotland/GCVR-170107/2020\|EPI_ISL_459696\|2 | ···················· | ···················· | ···················· | ···················· | ···················· | ···················· |
| C | 7088 hCoV-19/Scotland/GCVR-1702AA/2020\|EPI_ISL_459697\|2 | ···················· | ···················· | ···················· | ···················· | ···················· | ···················· |
| C | 7089 hCoV-19/Scotland/GCVR-1708B7/2020\|EPI_ISL_459699\|2 | ···················· | ·A·················· | ···················· | ···················· | ···················· | ···················· |
| C | 7090 hCoV-19/USA/MI-MDHHS-SC20552/2020\|EPI_ISL_460600\|2 | ···················· | ···················· | ···················· | ···················· | ···················· | ···················· |
| C | 7091 hCoV-19/USA/MI-MDHHS-SC20553/2020\|EPI_ISL_460601\|2 | ···················· | ···················· | ···················· | ···················· | ···················· | ···················· |
| C | 7092 hCoV-19/USA/MI-MDHHS-SC20554/2020\|EPI_ISL_460602\|2 | ···················· | ···················· | ···················· | ···················· | ·M·················· | ···················· |
| C | 7093 hCoV-19/Russia/CRIE145922/2020\|EPI_ISL_460604\|2020 | ···················· | ···················· | ···················· | ···················· | ···················· | ···················· |
| C | 7094 hCoV-19/Russia/CRIE140762/2020\|EPI_ISL_460605\|2020 | ···················· | ···················· | ···················· | ···················· | ···················· | ···················· |
| C | 7095 hCoV-19/Canada/BC_15926629/2020\|EPI_ISL_460609\|202 | ···················· | ···················· | ···················· | ···················· | ···················· | ···················· |
| C | 7096 hCoV-19/Netherlands/Friesland_28/2020\|EPI_ISL_4606 | ···················· | ···················· | ···················· | ···················· | ···················· | ···················· |
| C | 7097 hCoV-19/Netherlands/Gelderland_15/2020\|EPI_ISL_460 | ···················· | ···················· | ···················· | ···················· | ···················· | ···················· |
| C | 7098 hCoV-19/Netherlands/Gelderland_16/2020\|EPI_ISL_460 | ···················· | ···················· | ···················· | ···················· | ···················· | ···················· |
| C | 7099 hCoV-19/Netherlands/Gelderland_17/2020\|EPI_ISL_460 | ···················· | ···················· | ···················· | ···················· | ···················· | ···················· |
| C | 7100 hCoV-19/Netherlands/Gelderland_18/2020\|EPI_ISL_460 | ···················· | ···················· | ···················· | ···················· | ···················· | ···················· |
| C | 7101 hCoV-19/Netherlands/Gelderland_19/2020\|EPI_ISL_460 | ···················· | ···················· | ···················· | ···················· | ···················· | ···················· |
| C | 7102 hCoV-19/Netherlands/Gelderland_20/2020\|EPI_ISL_460 | ···················· | ···················· | ···················· | ···················· | ···················· | ···················· |
| C | 7103 hCoV-19/Netherlands/Gelderland_21/2020\|EPI_ISL_460 | ···················· | ···················· | ···················· | ···················· | ···················· | ···················· |
| C | 7104 hCoV-19/Netherlands/Gelderland_22/2020\|EPI_ISL_460 | ···················· | ···················· | ···················· | ···················· | ···················· | ···················· |
| C | 7105 hCoV-19/Netherlands/Gelderland_23/2020\|EPI_ISL_460 | ···················· | ···················· | ···················· | ···················· | ···················· | ···················· |
| C | 7106 hCoV-19/Netherlands/NA_487/2020\|EPI_ISL_460660\|202 | ···················· | ···················· | ···················· | ···················· | ···················· | ···················· |
| C | 7107 hCoV-19/Netherlands/NA_488/2020\|EPI_ISL_460661\|202 | ···················· | ···················· | ···················· | ···················· | ···················· | ···················· |
| C | 7108 hCoV-19/Netherlands/NA_489/2020\|EPI_ISL_460662\|202 | ···················· | ···················· | ···················· | ···················· | ···················· | ···················· |
| C | 7109 hCoV-19/Netherlands/NA_490/2020\|EPI_ISL_460663\|202 | ···················· | ···················· | ···················· | ···················· | ···················· | ···················· |
| C | 7110 hCoV-19/Netherlands/NA_491/2020\|EPI_ISL_460664\|202 | ···················· | ···················· | ···················· | ···················· | ···················· | ···················· |
| C | 7111 hCoV-19/Netherlands/NA_493/2020\|EPI_ISL_460666\|202 | ···················· | ···················· | ···················· | ···················· | ···················· | ···················· |
| C | 7112 hCoV-19/Netherlands/NA_494/2020\|EPI_ISL_460667\|202 | ···················· | ···················· | ···················· | ···················· | ···················· | ···················· |
| C | 7113 hCoV-19/Netherlands/NA_495/2020\|EPI_ISL_460668\|202 | ···················· | ···················· | ···················· | ···················· | ···················· | ···················· |
| C | 7114 hCoV-19/Netherlands/NA_496/2020\|EPI_ISL_460669\|202 | ···················· | ···················· | ···················· | ···················· | ···················· | ···················· |
| C | 7115 hCoV-19/Netherlands/NA_497/2020\|EPI_ISL_460670\|202 | ···················· | ···················· | ···················· | ···················· | ···················· | ···················· |
| C | 7116 hCoV-19/Netherlands/NA_498/2020\|EPI_ISL_460671\|202 | ···················· | ·A·················· | ···················· | ···················· | ···················· | ···················· |
| C | 7117 hCoV-19/Netherlands/NA_499/2020\|EPI_ISL_460672\|202 | ···················· | ···················· | ···················· | ···················· | ···················· | ···················· |
| C | 7118 hCoV-19/Netherlands/NA_500/2020\|EPI_ISL_460673\|202 | ···················· | ···················· | ···················· | ···················· | ···················· | ···················· |
| C | 7119 hCoV-19/Netherlands/NA_501/2020\|EPI_ISL_460674\|202 | ···················· | ···················· | ···················· | ···················· | ···················· | ···················· |
| C | 7120 hCoV-19/Netherlands/NA_502/2020\|EPI_ISL_460675\|202 | ···················· | ···················· | ···················· | ···················· | ···················· | ···················· |
| C | 7121 hCoV-19/Netherlands/NA_503/2020\|EPI_ISL_460676\|202 | ···················· | ···················· | ···················· | ···················· | ···················· | ···················· |
| C | 7122 hCoV-19/Netherlands/NA_504/2020\|EPI_ISL_460677\|202 | ···················· | ···················· | ···················· | ···················· | ···················· | ···················· |
| C | 7123 hCoV-19/Netherlands/NA_509/2020\|EPI_ISL_460682\|202 | ···················· | ···A················ | ···················· | ···················· | ···················· | ···················· |
| C | 7124 hCoV-19/Netherlands/NA_510/2020\|EPI_ISL_460683\|202 | ···················· | ···················· | ···················· | ···················· | ···················· | ···················· |
| C | 7125 hCoV-19/Netherlands/NA_511/2020\|EPI_ISL_460684\|202 | ···················· | ···················· | ···················· | ···················· | ···················· | ···················· |
| C | 7126 hCoV-19/Netherlands/NA_512/2020\|EPI_ISL_460685\|202 | ···················· | ···················· | ···················· | ···················· | ···················· | ···················· |
| C | 7127 hCoV-19/Netherlands/NA_513/2020\|EPI_ISL_460686\|202 | ···················· | ···················· | ···················· | ···················· | ···················· | ···················· |
| C | 7128 hCoV-19/Netherlands/NA_514/2020\|EPI_ISL_460687\|202 | ···················· | ···················· | ···················· | ···················· | ···················· | ···················· |
| C | 7129 hCoV-19/Netherlands/NA_515/2020\|EPI_ISL_460688\|202 | ···················· | ···················· | ···················· | ···················· | ···················· | ···················· |
| C | 7130 hCoV-19/Netherlands/NA_517/2020\|EPI_ISL_460690\|202 | ···················· | ···················· | ···················· | ···················· | ···················· | ···················· |
| C | 7131 hCoV-19/Netherlands/NA_518/2020\|EPI_ISL_460691\|202 | ···················· | ···················· | ···················· | ···················· | ···················· | ···················· |
| C | 7132 hCoV-19/Netherlands/NA_519/2020\|EPI_ISL_460692\|202 | ···················· | ···················· | ···················· | ···················· | ···················· | ···················· |
| C | 7133 hCoV-19/Netherlands/NA_520/2020\|EPI_ISL_460693\|202 | ···················· | ···················· | ···················· | ···················· | ···················· | ···················· |
| C | 7134 hCoV-19/Netherlands/NA_521/2020\|EPI_ISL_460694\|202 | ···················· | ···················· | ···················· | ···················· | ···················· | ···················· |
| C | 7135 hCoV-19/Netherlands/NA_522/2020\|EPI_ISL_460695\|202 | ···················· | ···················· | ···················· | ···················· | ···················· | ···················· |
| C | 7136 hCoV-19/Netherlands/NA_523/2020\|EPI_ISL_460696\|202 | ···················· | ···················· | ···················· | ···················· | ···················· | ···················· |
| C | 7137 hCoV-19/Netherlands/NA_524/2020\|EPI_ISL_460697\|202 | ···················· | ···················· | ···················· | ···················· | ···················· | ···················· |
| C | 7138 hCoV-19/Netherlands/NA_525/2020\|EPI_ISL_460698\|202 | ···················· | ···················· | ···················· | ···················· | ···················· | ···················· |
| C | 7139 hCoV-19/Netherlands/NA_526/2020\|EPI_ISL_460699\|202 | ···················· | ···················· | ···················· | ···················· | ···················· | ···················· |
| C | 7140 hCoV-19/England/NOTT-1116EB/2020\|EPI_ISL_461910\|20 | ···················· | ···················· | ···················· | ···················· | ···················· | ···················· |
| C | 7141 hCoV-19/England/NOTT-111751/2020\|EPI_ISL_461917\|20 | ···················· | ···················· | ···················· | ···················· | ···················· | ···················· |
| C | 7142 hCoV-19/England/NOTT-111760/2020\|EPI_ISL_461918\|20 | ···················· | ···················· | ···················· | ···················· | ···················· | ···················· |
| C | 7143 hCoV-19/England/NOTT-11177F/2020\|EPI_ISL_461919\|20 | ···················· | ···················· | ···················· | ···················· | ···················· | ···················· |
| C | 7144 hCoV-19/England/NOTT-11178E/2020\|EPI_ISL_461920\|20 | ···················· | ···················· | ···················· | ···················· | ···················· | ···················· |
| C | 7145 hCoV-19/England/NOTT-11179D/2020\|EPI_ISL_461921\|20 | ···················· | ···················· | ···················· | ···················· | ···················· | ···················· |
| C | 7146 hCoV-19/England/NOTT-1117AC/2020\|EPI_ISL_461922\|20 | ···················· | ···················· | ···················· | ···················· | ···················· | ···············A···· |
| C | 7147 hCoV-19/England/NOTT-1117BB/2020\|EPI_ISL_461923\|20 | ···················· | ···················· | ···················· | ···················· | ···················· | ···············A···· |
| C | 7148 hCoV-19/England/NOTT-1117CA/2020\|EPI_ISL_461924\|20 | ···················· | ···················· | ···················· | ···················· | ···················· | ···················· |
| C | 7149 hCoV-19/England/NOTT-1117D9/2020\|EPI_ISL_461925\|20 | ···················· | ···················· | ···················· | ···················· | ···················· | ···················· |
| C | 7150 hCoV-19/England/NOTT-1117E8/2020\|EPI_ISL_461926\|20 | ···················· | ···················· | ···················· | ···················· | ···················· | ···················· |
| C | 7151 hCoV-19/England/NOTT-1117F7/2020\|EPI_ISL_461927\|20 | ···················· | ···················· | ···················· | ···················· | ···················· | ···················· |
| C | 7152 hCoV-19/England/NOTT-111803/2020\|EPI_ISL_461928\|20 | ···················· | ···················· | ···················· | ···················· | ···················· | ···················· |
| C | 7153 hCoV-19/England/NOTT-111821/2020\|EPI_ISL_461929\|20 | ···················· | ···················· | ···················· | ···················· | ···················· | ···················· |
| C | 7154 hCoV-19/England/NOTT-111830/2020\|EPI_ISL_461930\|20 | ···················· | ···················· | ···················· | ···················· | ···················· | ···················· |
| C | 7155 hCoV-19/England/NOTT-11184F/2020\|EPI_ISL_461931\|20 | ···················· | ···················· | ···················· | ···················· | ···················· | ···················· |
| C | 7156 hCoV-19/England/NOTT-11185E/2020\|EPI_ISL_461932\|20 | ···················· | ···················· | ···················· | ···················· | ···················· | ···················· |
| C | 7157 hCoV-19/England/NOTT-11188B/2020\|EPI_ISL_461934\|20 | ···················· | ···················· | ···················· | ···················· | ···················· | ···················· |
| C | 7158 hCoV-19/England/NOTT-11189A/2020\|EPI_ISL_461935\|20 | ···················· | ···················· | ···················· | ···················· | ···················· | ···················· |
| C | 7159 hCoV-19/England/NOTT-1118A9/2020\|EPI_ISL_461936\|20 | ···················· | ···················· | ···················· | ···················· | ···················· | ···················· |
| C | 7160 hCoV-19/England/NOTT-1118B8/2020\|EPI_ISL_461937\|20 | ···················· | ···················· | ···················· | ···················· | ···················· | ···················· |
| C | 7161 hCoV-19/England/PORT-2CF78E/2020\|EPI_ISL_461981\|20 | ···················· | ···················· | ···················· | ···················· | ···················· | ···················· |
| C | 7162 hCoV-19/England/PORT-2CFC8F/2020\|EPI_ISL_461983\|20 | ···················· | ···················· | ···················· | ···················· | ···················· | ···················· |
| C | 7163 hCoV-19/England/PORT-2CFCCB/2020\|EPI_ISL_461986\|20 | ···················· | ···················· | ···················· | ···················· | ···················· | ···················· |
| C | 7164 hCoV-19/England/PORT-2CFCDA/2020\|EPI_ISL_461987\|20 | ···················· | ···················· | ···················· | ···················· | ···················· | ···················· |

| | | | | | | | | |
|---|---|---|---|---|---|---|---|---|
| C | 7165 hCoV-19/England/PORT-2CFD13/2020\|EPI_ISL_461989\|20 | ···················· | ···················· | ···················· | ···················· | ···················· | ···················· | |
| C | 7166 hCoV-19/England/PORT-2CFD40/2020\|EPI_ISL_461991\|20 | ···················· | ···················· | ···················· | ···················· | ···················· | ···················· | |
| C | 7167 hCoV-19/England/PORT-2CFDB9/2020\|EPI_ISL_461998\|20 | ···················· | ···················· | ···················· | ···················· | ···················· | ···················· | |
| C | 7168 hCoV-19/England/SHEF-C0CF9/2020\|EPI_ISL_461999\|202 | ···················· | ···················· | ···················· | ···················· | ···················· | ···················· | |
| C | 7169 hCoV-19/DRC/3595/2020\|EPI_ISL_447606\|2020-04-23 | ···················· | ···················· | ···················· | ···················· | ···················· | ···················· | |
| C | 7170 hCoV-19/DRC/3632/2020\|EPI_ISL_447607\|2020-04-23 | ···················· | ···················· | ···················· | ···················· | ···················· | ···················· | |
| C | 7171 hCoV-19/Taiwan/NTU10/2020\|EPI_ISL_447614\|2020-03-1 | ···················· | ···················· | ···················· | ···················· | ···················· | ···················· | |
| C | 7172 hCoV-19/Taiwan/NTU18/2020\|EPI_ISL_447615\|2020-03-1 | ···················· | ···················· | ···················· | ···················· | ···················· | ···················· | |
| C | 7173 hCoV-19/Taiwan/NTU22/2020\|EPI_ISL_447616\|2020-03-2 | ···················· | ···················· | ···················· | ···················· | ···················· | ···················· | |
| C | 7174 hCoV-19/Taiwan/NTU23/2020\|EPI_ISL_447617\|2020-03-2 | ···················· | ···················· | ···················· | ···················· | ···················· | ···················· | |
| C | 7175 hCoV-19/Taiwan/NTU24/2020\|EPI_ISL_447618\|2020-03-2 | ···················· | ···················· | ···················· | ···················· | ···················· | ···················· | |
| C | 7176 hCoV-19/Taiwan/NTU25/2020\|EPI_ISL_447619\|2020-03-2 | ···················· | ···················· | ···················· | ···················· | ···················· | ···················· | |
| C | 7177 hCoV-19/Taiwan/NTU26/2020\|EPI_ISL_447620\|2020-03-3 | ···················· | ···················· | ···················· | ···················· | ···················· | ···················· | |
| C | 7178 hCoV-19/Taiwan/NTU27/2020\|EPI_ISL_447621\|2020-04-1 | ···················· | ···················· | ···················· | ···················· | ···················· | ···················· | |
| C | 7179 hCoV-19/Taiwan/NTU28/2020\|EPI_ISL_447622\|2020-04-2 | ···················· | ···················· | ···················· | ···················· | ···················· | ···················· | |
| C | 7180 hCoV-19/Northern Ireland/NIRE-FAF75/2020\|EPI_ISL_4 | ···················· | ···················· | ···················· | ···················· | ···················· | ···················· | |
| C | 7182 hCoV-19/Northern Ireland/NIRE-FB1D8/2020\|EPI_ISL_4 | ···················· | ···················· | ···················· | ···················· | ···················· | ···················· | |
| C | 7184 hCoV-19/Northern Ireland/NIRE-FB220/2020\|EPI_ISL_4 | ···················· | ···················· | ···················· | ···················· | ···················· | ···················· | |
| C | 7186 hCoV-19/England/NORW-E6DDC/2020\|EPI_ISL_448979\|202 | ···················· | ···················· | ···················· | ···················· | ···················· | ···················· | |
| C | 7187 hCoV-19/England/NORW-E6DEB/2020\|EPI_ISL_448980\|202 | ···················· | ···················· | ···················· | ···················· | ···················· | ···················· | |
| C | 7188 hCoV-19/England/NORW-E6E15/2020\|EPI_ISL_448981\|202 | ···················· | ···················· | ···················· | ···················· | ···················· | ···················· | |
| C | 7189 hCoV-19/England/NORW-E6E24/2020\|EPI_ISL_448982\|202 | ···················· | ···················· | ···················· | ···················· | ···················· | ···················· | |
| C | 7190 hCoV-19/England/NORW-E6E42/2020\|EPI_ISL_448983\|202 | ···················· | ···················· | ···················· | ···················· | ···················· | ···················· | |
| C | 7191 hCoV-19/England/NORW-E6E60/2020\|EPI_ISL_448984\|202 | ···················· | ···················· | ···················· | ···················· | ···················· | ···················· | |
| C | 7192 hCoV-19/England/NORW-E6E8E/2020\|EPI_ISL_448985\|202 | ···················· | ···················· | ··········Y········· | ···················· | ···················· | ···················· | |
| C | 7193 hCoV-19/England/NORW-E6E9D/2020\|EPI_ISL_448986\|202 | ···················· | ···················· | ···················· | ···················· | ···················· | ···················· | |
| C | 7194 hCoV-19/England/NORW-E6EAC/2020\|EPI_ISL_448987\|202 | ···················· | ···················· | ···················· | ···················· | ···················· | ···················· | |
| C | 7195 hCoV-19/England/NORW-E6EBB/2020\|EPI_ISL_448988\|202 | ···················· | ···················· | ···················· | ···················· | ···················· | ···················· | |
| C | 7196 hCoV-19/England/NORW-E6F03/2020\|EPI_ISL_448991\|202 | ···················· | ···················· | ···················· | ···················· | NNN················· | ···················· | 1 |
| C | 7197 hCoV-19/England/NORW-E6F12/2020\|EPI_ISL_448992\|202 | ···················· | ···················· | ···················· | ···················· | ···················· | ···················· | |
| C | 7198 hCoV-19/England/NORW-E6F21/2020\|EPI_ISL_448993\|202 | ···················· | ···················· | ···················· | ···················· | ···················· | ···················· | |
| C | 7200 hCoV-19/England/NORW-E6FA9/2020\|EPI_ISL_448995\|202 | ···················· | ···················· | ···················· | ···················· | ···················· | ···················· | |
| C | 7201 hCoV-19/England/NORW-E6FC7/2020\|EPI_ISL_448996\|202 | ···················· | ···················· | ···················· | ···················· | ···················· | ···················· | |
| C | 7203 hCoV-19/England/NORW-E705A/2020\|EPI_ISL_448999\|202 | ···················· | ···················· | ···················· | ···················· | ···················· | ···················· | |
| C | 7204 hCoV-19/Scotland/CVR3153/2020\|EPI_ISL_449190\|2020- | ···················· | ···················A | ···················· | ···················· | ···················· | ···················· | 1 |
| C | 7205 hCoV-19/Scotland/CVR3154/2020\|EPI_ISL_449191\|2020- | ···················· | ···················· | ···················· | ···················· | ···················· | ···················· | |
| C | 7206 hCoV-19/Scotland/CVR3156/2020\|EPI_ISL_449192\|2020- | ···················· | ···················A | ···················· | ···················· | ···················· | ···················· | 1 |
| C | 7207 hCoV-19/Scotland/CVR3157/2020\|EPI_ISL_449193\|2020- | ···················· | ···················· | ···················· | ···················· | ···················· | ···················· | |
| C | 7208 hCoV-19/Scotland/CVR3158/2020\|EPI_ISL_449194\|2020- | ···················· | ···················· | ···················· | ···················· | ···················· | ···················· | |
| C | 7209 hCoV-19/Scotland/CVR3160/2020\|EPI_ISL_449195\|2020- | ···················· | ···················A | ···················· | ···················· | ···················· | ···················· | 1 |
| C | 7210 hCoV-19/Scotland/CVR3162/2020\|EPI_ISL_449196\|2020- | ···················· | ···················A | ···················· | ···················· | ···················· | ···················· | 1 |
| C | 7211 hCoV-19/Scotland/CVR3163/2020\|EPI_ISL_449197\|2020- | ···················· | ···················· | ···················· | ···················· | ···················· | ···················· | |
| C | 7212 hCoV-19/Scotland/CVR3165/2020\|EPI_ISL_449198\|2020- | ···················· | ···················· | ···················· | ···················· | ···················· | ···················· | |
| C | 7213 hCoV-19/Scotland/CVR3169/2020\|EPI_ISL_449199\|2020- | ···················· | ···················· | ···················· | ···················· | ···················· | ···················· | |
| C | 7214 hCoV-19/USA/LA-SR0323/2020\|EPI_ISL_450177\|2020-04- | ···················· | ···················· | ···················· | ···················· | ···················· | ···················· | |
| C | 7215 hCoV-19/USA/LA-SR0324/2020\|EPI_ISL_450178\|2020-04- | ···················· | ···················· | ···················· | ···················· | ···················· | ···················· | |
| C | 7216 hCoV-19/USA/LA-SR0326/2020\|EPI_ISL_450179\|2020-04- | ···················· | ···················· | ···················· | ···················· | ···················· | ···················· | |
| C | 7217 hCoV-19/USA/LA-SR0327/2020\|EPI_ISL_450180\|2020-04- | ···················· | ···················· | ···················· | ···················· | ···················· | ···················· | |
| C | 7218 hCoV-19/USA/LA-SR0328/2020\|EPI_ISL_450181\|2020-04- | ···················· | ···················· | ···················· | ···················· | ···················· | ···················· | |
| C | 7219 hCoV-19/USA/LA-SR0329/2020\|EPI_ISL_450182\|2020-04- | ···················· | ···················· | ···················· | ···················· | ···················· | ···················· | |
| C | 7220 hCoV-19/USA/LA-SR0331/2020\|EPI_ISL_450184\|2020-04- | ···················· | ···················· | ···················· | ···················· | ···················· | ···················· | |
| C | 7221 hCoV-19/USA/LA-SR0332/2020\|EPI_ISL_450185\|2020-04- | ···················· | ···················· | ···················· | ···················· | ···················· | ···················· | |
| C | 7222 hCoV-19/Jordan/SR-0333/2020\|EPI_ISL_450186\|2020-04 | ···················· | ·············A······ | ···················· | ···················· | ···················· | ···················· | |
| C | 7223 hCoV-19/Jordan/SR-0335/2020\|EPI_ISL_450187\|2020-04 | ···················· | ···················· | ···················· | ···················· | ···················· | ···················· | |
| C | 7224 hCoV-19/Jordan/SR-0336/2020\|EPI_ISL_450188\|2020-04 | ···················· | ···················· | ···················· | ···················· | ···················· | ···················· | |
| C | 7225 hCoV-19/USA/CA-SR0341/2020\|EPI_ISL_450190\|2020-05- | ···················· | ·A·················· | ···················· | ···················· | ···················· | ···················· | |
| C | 7226 hCoV-19/USA/CA-SR0347/2020\|EPI_ISL_450191\|2020-03- | ···················· | ···················· | ···················· | ···················· | ···················· | ···················· | |
| C | 7227 hCoV-19/Australia/SAP029/2020\|EPI_ISL_450193\|2020- | ···················· | ···················· | ···················· | ···················· | ···················· | ···················· | |
| C | 7228 hCoV-19/Australia/SAP036/2020\|EPI_ISL_450194\|2020- | ···················· | ···················· | ···················· | ···················· | ···················· | ···················· | |
| C | 7229 hCoV-19/Thailand/Bangkok_168/2020\|EPI_ISL_450196\|2 | ···················· | ···················· | ···················· | ···················· | ···················· | ···················· | |
| C | 7230 hCoV-19/Thailand/Nonthaburi_2520/2020\|EPI_ISL_4501 | ···················· | ···················· | ···················· | ···················· | ···················· | ···················· | |
| C | 7231 hCoV-19/New Zealand/20VR1903/2020\|EPI_ISL_456302\|2 | ···················· | ···················· | ···················· | ···················· | ···················· | ···················· | |
| C | 7232 hCoV-19/New Zealand/20VR1907/2020\|EPI_ISL_456303\|2 | ···················· | ···················· | ···················· | ···················· | ···················· | ···················· | |
| C | 7233 hCoV-19/New Zealand/20VR1908/2020\|EPI_ISL_456304\|2 | ···················· | ···················· | ···················· | ···················· | ···················· | ···················· | |
| C | 7234 hCoV-19/New Zealand/20VR1910/2020\|EPI_ISL_456305\|2 | ···················· | ···················· | ···················· | ···················· | ···················· | ···················· | |
| C | 7235 hCoV-19/New Zealand/20VR1917/2020\|EPI_ISL_456308\|2 | ···················· | ···················· | ···················· | ···················· | ···················· | ···················· | |
| C | 7236 hCoV-19/New Zealand/20VR1924/2020\|EPI_ISL_456309\|2 | ···················· | ···················· | ···················· | ···················· | ···················· | ···················· | |
| C | 7237 hCoV-19/New Zealand/20VR1925/2020\|EPI_ISL_456310\|2 | ···················· | ···················· | ···················· | ···················· | ···················· | ···················· | |
| C | 7238 hCoV-19/New Zealand/20VR1933/2020\|EPI_ISL_456314\|2 | ···················· | ···················· | ···················· | ···················· | ···················· | ···················· | |
| C | 7240 hCoV-19/Scotland/EDB5344/2020\|EPI_ISL_457002\|2020- | ···················· | ·A·················· | ···················· | ···················· | ···················· | ···················· | |
| C | 7241 hCoV-19/Scotland/EDB5345/2020\|EPI_ISL_457003\|2020- | ···················· | ·A·················· | ···················· | ···················· | ···················· | ···················· | |
| C | 7242 hCoV-19/Scotland/EDB5348/2020\|EPI_ISL_457004\|2020- | ···················· | ·A·················· | ···················· | ···················· | ···················· | ···················· | |
| C | 7243 hCoV-19/Scotland/EDB5349/2020\|EPI_ISL_457005\|2020- | ···················· | ···················· | ···················· | ···················· | ···················· | ···················· | |
| C | 7244 hCoV-19/Scotland/EDB5351/2020\|EPI_ISL_457006\|2020- | ···················· | ·A·················· | ···················· | ···················· | ···················· | ···················· | |
| C | 7245 hCoV-19/England/SHEF-C7EDE/2020\|EPI_ISL_457624\|202 | ···················· | ···················· | ···················· | ···················· | ···················· | ···················· | |
| C | 7246 hCoV-19/England/SHEF-C8514/2020\|EPI_ISL_457627\|202 | ···················· | ···················· | ···················· | ···················· | ···················· | ···················· | |
| C | 7247 hCoV-19/England/SHEF-C8A33/2020\|EPI_ISL_457633\|202 | ···················· | ···················· | ···················· | ···················· | ···················· | ···················· | |
| C | 7248 hCoV-19/England/SHEF-C930A/2020\|EPI_ISL_457634\|202 | ···················· | ···················· | ···················· | ···················· | ···················· | ···················· | |

C 7249 hCoV-19/England/SHEF-C95B9/2020|EPI_ISL_457636|202 ···················· ···················· ···················· ···················· ···················· ····················
C 7250 hCoV-19/England/SHEF-C9A05/2020|EPI_ISL_457637|202 ···················· ···················· ···················· ···················· ···················· ····················
C 7251 hCoV-19/England/SHEF-CA266/2020|EPI_ISL_457640|202 ···················· ···················· ···················· ···················· ···················· ····················
C 7252 hCoV-19/England/SHEF-CA354/2020|EPI_ISL_457641|202 ···················· ···················· ···················· ···················· ···················· ····················
C 7253 hCoV-19/England/SHEF-CAA5F/2020|EPI_ISL_457643|202 ···················· ···················· ···················· ···················· ···················· ····················
C 7254 hCoV-19/England/BRIS-12A676/2020|EPI_ISL_458300|20 ···················· ···················· ···················· ···················· ···················· ····················
C 7255 hCoV-19/England/BRIS-12A843/2020|EPI_ISL_458301|20 ···················· ···················· ···················· ···················· ···················· ····················
C 7256 hCoV-19/England/BRIS-129862/2020|EPI_ISL_458302|20 ···················· ···················· ···················· ···················· ···················· ····················
C 7257 hCoV-19/England/BRIS-12A2BE/2020|EPI_ISL_458303|20 ···················· ···················· ···················· ···················· ···················· ····················
C 7258 hCoV-19/England/BRIS-12A719/2020|EPI_ISL_458304|20 ···················· ···················· ···················· ···················· ···················· ····················
C 7259 hCoV-19/England/BRIS-129E33/2020|EPI_ISL_458305|20 ···················· ···················· ···················· ···················· ···················· ····················
C 7260 hCoV-19/England/BRIS-12A245/2020|EPI_ISL_458306|20 ···················· ···················· ···················· ···················· ···················· ····················
C 7261 hCoV-19/England/BRIS-12A37F/2020|EPI_ISL_458308|20 ···················· ···················· ···················· ···················· ···················· ····················
C 7262 hCoV-19/England/BRIS-129DDC/2020|EPI_ISL_458311|20 ···················· ···················· ···················· ···················· ···················· ····················
C 7263 hCoV-19/England/BRIS-12AC1A/2020|EPI_ISL_458313|20 ···················· ···················· ···················· ···················· ···················· ····················
C 7264 hCoV-19/England/BRIS-12A227/2020|EPI_ISL_458316|20 ···················· ···················· ···················· ···················· ···················· ····················
C 7265 hCoV-19/England/BRIS-129EBB/2020|EPI_ISL_458317|20 ···················· ···················· ···················· ···················· ···················· ····················
C 7266 hCoV-19/England/BRIS-12AED8/2020|EPI_ISL_458319|20 ···················· ···················· ···················· ···················· ···················· ····················
C 7267 hCoV-19/England/BRIS-129A99/2020|EPI_ISL_458320|20 ···················· ···················· ···················· ···················· ···················· ····················
C 7268 hCoV-19/England/BRIS-12A658/2020|EPI_ISL_458321|20 ···················· ···················· ···················· ···················· ···················· ····················
C 7269 hCoV-19/England/BRIS-12ABA4/2020|EPI_ISL_458323|20 ···················· ···················· ···················· ···················· ···················· ····················
C 7270 hCoV-19/England/BRIS-12A3AC/2020|EPI_ISL_458324|20 ···················· ···················· ···················· ···················· ···················· ····················
C 7271 hCoV-19/England/BRIS-12AC65/2020|EPI_ISL_458329|20 ···················· ···················· ···················· ···················· ···················· ····················
C 7272 hCoV-19/England/BRIS-12AD9F/2020|EPI_ISL_458330|20 ···················· ···················· ···················· ···················· ···················· ····················
C 7273 hCoV-19/England/BRIS-12A290/2020|EPI_ISL_458332|20 ···················· ···················· ···················· ···················· ···················· ····················
C 7274 hCoV-19/England/BRIS-12ADCC/2020|EPI_ISL_458333|20 ···················· ···················· ···················· ···················· ···················· ····················
C 7275 hCoV-19/England/BRIS-12A649/2020|EPI_ISL_458334|20 ···················· ···················· ···················· ···················· ···················· ····················
C 7276 hCoV-19/England/BRIS-12AB77/2020|EPI_ISL_458337|20 ···················· ···················· ···················· ···················· ···················· ····················
C 7277 hCoV-19/England/BRIS-12A98C/2020|EPI_ISL_458338|20 ···················· ···················· ···················· ···················· ···················· ····················
C 7278 hCoV-19/England/BRIS-128DCE/2020|EPI_ISL_458903|20 ···················· ···················· ···················· ···················· ···················· ····················
C 7279 hCoV-19/England/BRIS-12971A/2020|EPI_ISL_458904|20 ···················· ···················· ···················· ···················· ···················· ····················
C 7280 hCoV-19/England/BRIS-12961D/2020|EPI_ISL_458908|20 ···················· ···················· ···················· ···················· ···················· ····················
C 7281 hCoV-19/England/BRIS-128A8B/2020|EPI_ISL_458909|20 ···················· ···················· ···················· ···················· ···················· ····················
C 7282 hCoV-19/England/BRIS-129325/2020|EPI_ISL_458911|20 ···················· ···················· ···················· ···················· ···················· ····················
C 7283 hCoV-19/England/BRIS-126373/2020|EPI_ISL_458912|20 ···················· ···················· ···················· ···················· ···················· ····················
C 7284 hCoV-19/England/BRIS-1266A7/2020|EPI_ISL_458913|20 ···················· ···················· ···················· ···················· ···················· ····················
C 7285 hCoV-19/England/BRIS-1269CC/2020|EPI_ISL_458914|20 ···················· ···················· ···················· ···················· ···················· ····················
C 7286 hCoV-19/England/BRIS-126B11/2020|EPI_ISL_458915|20 ···················· ···················· ···················· ···················· ···················· ····················
C 7287 hCoV-19/England/BRIS-1261C4/2020|EPI_ISL_458917|20 ···················· ·············A······ ···················· ···················· ···················· ····················
C 7288 hCoV-19/England/BRIS-126C78/2020|EPI_ISL_458918|20 ···················· ···················· ···················· ···················· ···················· ····················
C 7289 hCoV-19/England/BRIS-126CC3/2020|EPI_ISL_458919|20 ···················· ···················· ···················· ···················· ···················· ····················
C 7290 hCoV-19/England/BRIS-12612E/2020|EPI_ISL_458922|20 ···················· ···················· ···················· ···················· ···················· ····················
C 7291 hCoV-19/England/BRIS-126258/2020|EPI_ISL_458923|20 ···················· ···················· ···················· ···················· ···················· ····················
C 7292 hCoV-19/England/BRIS-126030/2020|EPI_ISL_458924|20 ···················· ···················· ···················· ···················· ···················· ····················
C 7293 hCoV-19/England/BRIS-126407/2020|EPI_ISL_458925|20 ···················· ···················· ···················· ···················· ···················· ····················
C 7294 hCoV-19/England/BRIS-12630A/2020|EPI_ISL_458926|20 ···················· ···················· ···················· ···················· ···················· ····················
C 7295 hCoV-19/England/BRIS-12605E/2020|EPI_ISL_458927|20 ···················· ···················· ···················· ···················· ···················· ····················
C 7296 hCoV-19/England/BRIS-12615B/2020|EPI_ISL_458928|20 ···················· ···················· ···················· ···················· ···················· ····················
C 7297 hCoV-19/England/BRIS-1264CB/2020|EPI_ISL_458930|20 ···················· ···················· ···················· ···················· ···················· ····················
C 7298 hCoV-19/England/BRIS-126461/2020|EPI_ISL_458931|20 ···················· ·A·················· ···················· ···················· ···················· ····················
C 7299 hCoV-19/England/BRIS-126A23/2020|EPI_ISL_458932|20 ···················· ···················· ···················· ···················· ···················· ····················
C 7300 hCoV-19/England/BRIS-1268CF/2020|EPI_ISL_458955|20 ···················· ···················· ···················· ···················· ···················· ····················
C 7301 hCoV-19/England/BRIS-1269BD/2020|EPI_ISL_458956|20 ···················· ····A··············· ···················· ···················· ···················· ····················
C 7302 hCoV-19/England/BRIS-12623A/2020|EPI_ISL_458957|20 ···················· ···················· ···················· ···················· ···················· ····················
C 7303 hCoV-19/England/BRIS-126CA5/2020|EPI_ISL_458960|20 ···················· ···················· ···················· ···················· ···················· ····················
C 7304 hCoV-19/England/BRIS-126AAB/2020|EPI_ISL_458961|20 ···················· ···················· ···················· ···················· ···················· ····················
C 7305 hCoV-19/England/BRIS-126DA2/2020|EPI_ISL_458964|20 ···················· ···················· ···················· ···················· ···················· ····················
C 7306 hCoV-19/England/BRIS-1264BC/2020|EPI_ISL_458965|20 ···················· ···················· ···················· ···················· ···················· ····················
C 7307 hCoV-19/England/BRIS-126364/2020|EPI_ISL_458966|20 ···················· ···················· ···················· ···················· ···················· ····················
C 7308 hCoV-19/England/BRIS-126DEE/2020|EPI_ISL_458967|20 ···················· ···················· ···················· ···················· ···················· ····················
C 7309 hCoV-19/England/BRIS-126829/2020|EPI_ISL_458969|20 ···················· ···················· ···················· ···················· ···················· ····················
C 7310 hCoV-19/England/BRIS-12667A/2020|EPI_ISL_458971|20 ···················· ···················· ···················· ···················· ···················· ····················
C 7311 hCoV-19/England/BRIS-1267C2/2020|EPI_ISL_458972|20 ···················· ···················· ···················· ···················· ···················· ····················
C 7312 hCoV-19/England/BRIS-1267B3/2020|EPI_ISL_458974|20 ···················· ···················· ···················· ···················· ···················· ····················
C 7313 hCoV-19/England/BRIS-126B3F/2020|EPI_ISL_458975|20 ···················· ···················· ···················· ···················· ···················· ····················
C 7314 hCoV-19/England/BRIS-126416/2020|EPI_ISL_458976|20 ···················· ···················· ···················· ···················· ···················· ····················
C 7315 hCoV-19/Scotland/GCVR-1702C8/2020|EPI_ISL_459626|2 ···················· ···················· ···················· ···················· ···················· ····················
C 7316 hCoV-19/Scotland/GCVR-170705/2020|EPI_ISL_459628|2 ···················· ···················· ···················· ···················· ···················· ····················
C 7317 hCoV-19/Scotland/GCVR-170FA3/2020|EPI_ISL_459629|2 ···················· ···················· ···················· ···················· ···················· ····················
C 7318 hCoV-19/Scotland/GCVR-16FF89/2020|EPI_ISL_459630|2 ···················· ···················· ···················· ···················· ···················· ····················
C 7319 hCoV-19/Scotland/GCVR-1709D2/2020|EPI_ISL_459631|2 ···················· ···················A ···················· ···················· ···················· ···················· 1
C 7320 hCoV-19/Scotland/GCVR-17086C/2020|EPI_ISL_459632|2 ···················· ···················A ···················· ···················· ···················· ···················· 1
C 7321 hCoV-19/Scotland/GCVR-1705FC/2020|EPI_ISL_459635|2 ···················· ···················· ···················· ···················· ···················· ····················
C 7322 hCoV-19/Scotland/GCVR-170495/2020|EPI_ISL_459636|2 ···················· ···················· ···················· ···················· ···················· ····················
C 7323 hCoV-19/Scotland/GCVR-1704E0/2020|EPI_ISL_459637|2 ···················· ·A·················· ···················· ···················· ···················· ····················
C 7324 hCoV-19/Scotland/GCVR-17128B/2020|EPI_ISL_459638|2 ···················· ···················· ···················· ···················· ···················· ····················
C 7325 hCoV-19/Scotland/GCVR-170608/2020|EPI_ISL_459640|2 ···················· ···················· ···················· ···················· ···················· ····················
C 7326 hCoV-19/Scotland/GCVR-170F58/2020|EPI_ISL_459641|2 ···················· ···················· ···················· ···················· ···················· ····················

| | | | | | | | | |
|---|---|---|---|---|---|---|---|---|
| C | 7327 hCoV-19/Scotland/GCVR-17134C/2020\|EPI_ISL_459643\|2 | ···················· | ···················· | ···················· | ···················· | ···················· | ···················· | |
| C | 7328 hCoV-19/Scotland/GCVR-170398/2020\|EPI_ISL_459644\|2 | ···················· | ·A·················· | ···················· | ···················· | ···················· | ···················· | |
| C | 7329 hCoV-19/Scotland/GCVR-1702E6/2020\|EPI_ISL_459645\|2 | ···················· | ···················· | ···················· | ···················· | ···················· | ···················· | |
| C | 7330 hCoV-19/Scotland/GCVR-1703E3/2020\|EPI_ISL_459646\|2 | ···················· | ···················A | ···················· | ···················· | ···················· | ···················· | 1 |
| C | 7331 hCoV-19/Scotland/GCVR-170644/2020\|EPI_ISL_459647\|2 | ···················· | ···················· | ···················· | ···················· | ···················· | ···················· | |
| C | 7332 hCoV-19/Scotland/GCVR-170143/2020\|EPI_ISL_459648\|2 | ···················· | ···················· | ···················· | ···················· | ···················· | ···················· | |
| C | 7333 hCoV-19/Scotland/GCVR-170F67/2020\|EPI_ISL_459649\|2 | ···················· | ···················· | ···················· | ···················· | ···················· | ···················· | |
| C | 7334 hCoV-19/Scotland/GCVR-1700FB/2020\|EPI_ISL_459651\|2 | ···················· | ···················· | ···················· | ···················· | ···················· | ···················· | |
| C | 7335 hCoV-19/Scotland/GCVR-170653/2020\|EPI_ISL_459652\|2 | ···················· | ···················· | ···················· | ···················· | ···················· | ···················· | |
| C | 7336 hCoV-19/Scotland/GCVR-170CCA/2020\|EPI_ISL_459654\|2 | ···················· | ···················· | ···················· | ···················· | ···················· | ···················· | |
| C | 7337 hCoV-19/Scotland/GCVR-171054/2020\|EPI_ISL_459656\|2 | ···················· | ···················· | ···················· | ···················· | ···················· | ···················· | |
| C | 7338 hCoV-19/Scotland/GCVR-170B09/2020\|EPI_ISL_459657\|2 | ···················· | ···················· | ···················· | ···················· | ···················· | ···················· | |
| C | 7339 hCoV-19/Scotland/GCVR-170A48/2020\|EPI_ISL_459659\|2 | ···················· | ···················· | ···················· | ···················· | ···················· | ···················· | |
| C | 7340 hCoV-19/Scotland/GCVR-1709B4/2020\|EPI_ISL_459660\|2 | ···················· | ···················A | ···················· | ···················· | ···················· | ···················· | 1 |
| C | 7341 hCoV-19/Scotland/GCVR-170978/2020\|EPI_ISL_459661\|2 | ···················· | ···················· | ···················· | ···················· | ···················· | ···················· | |
| C | 7342 hCoV-19/Scotland/GCVR-1709A5/2020\|EPI_ISL_459662\|2 | ···················· | ···················· | ···················· | ···················· | ···················· | ···················· | |
| C | 7343 hCoV-19/Scotland/GCVR-170A0C/2020\|EPI_ISL_459665\|2 | ···················· | ···················· | ···················· | ···················· | ···················· | ···················· | |
| C | 7344 hCoV-19/Scotland/GCVR-1710EB/2020\|EPI_ISL_459666\|2 | ···················· | ···················· | ···················· | ···················· | ···················· | ···················· | |
| C | 7345 hCoV-19/Scotland/GCVR-17050B/2020\|EPI_ISL_459667\|2 | ···················· | ···················· | ···················· | ···················· | ···················· | ···················· | |
| C | 7346 hCoV-19/Scotland/GCVR-170B90/2020\|EPI_ISL_459668\|2 | ···················· | ···················A | ···················· | ···················· | ···················· | ···················· | 1 |
| C | 7347 hCoV-19/Canada/BC_16344159/2020\|EPI_ISL_460610\|202 | ···················· | ···················· | ···················· | ···················· | ···················· | ···················· | |
| C | 7348 hCoV-19/Canada/BC_17045358/2020\|EPI_ISL_460611\|202 | ···················· | ···················· | ···················· | ···················· | ···················· | ···················· | |
| C | 7349 hCoV-19/Canada/BC_17422770/2020\|EPI_ISL_460612\|202 | ···················· | ···················· | ···················· | ···················· | ···················· | ···················· | |
| C | 7350 hCoV-19/Canada/BC_17548428/2020\|EPI_ISL_460614\|202 | ···················· | ···················· | ···················· | ···················· | ···················· | ···················· | |
| C | 7351 hCoV-19/Canada/BC_18097749/2020\|EPI_ISL_460615\|202 | ···················· | ···················· | ···················· | ···················· | ···················· | ···················· | |
| C | 7352 hCoV-19/Canada/BC_18111443/2020\|EPI_ISL_460616\|202 | ···················· | ···················· | ···················· | ···················· | ···················· | ···················· | |
| C | 7353 hCoV-19/Turkey/IMU-SP-02/2020\|EPI_ISL_460617\|2020- | ···················· | ···················· | ···················· | ···················· | ···················· | ···················· | |
| C | 7354 hCoV-19/Turkey/IMU-SP-01/2020\|EPI_ISL_460618\|2020- | ···················· | ···················· | ···················· | ···················· | ···················· | ···················· | |
| C | 7355 hCoV-19/Turkey/IMU-SP-04/2020\|EPI_ISL_460619\|2020- | ···················· | ···················· | ···················· | ···················· | ···················· | ···················· | |
| C | 7356 hCoV-19/USA/WA-UW-4545/2020\|EPI_ISL_460621\|2020-03 | ···················· | ···················· | ···················· | ···················· | ···················· | ···················· | |
| C | 7357 hCoV-19/USA/ID-UW-4550/2020\|EPI_ISL_460622\|2020-03 | ···················· | ···················· | ···················· | ···················· | ···················· | ···················· | |
| C | 7358 hCoV-19/USA/WA-UW-4555/2020\|EPI_ISL_460623\|2020-03 | ···················· | ···················· | ···················· | ···················· | ···················· | ···················· | |
| C | 7359 hCoV-19/USA/WA-UW-4562/2020\|EPI_ISL_460624\|2020-03 | ···················· | ···················· | ···················· | ···················· | ···················· | ···················· | |
| C | 7360 hCoV-19/USA/WA-UW-4570/2020\|EPI_ISL_460625\|2020-03 | ···················· | ···················· | ···················· | ···················· | ···················· | ···················· | |
| C | 7361 hCoV-19/USA/WA-UW-4588/2020\|EPI_ISL_460626\|2020-03 | ···················· | ···················· | ···················· | ···················· | ···················· | ···················· | |
| C | 7362 hCoV-19/USA/WA-UW-4591/2020\|EPI_ISL_460627\|2020-03 | ···················· | ···················· | ···················· | ···················· | ···················· | ···················· | |
| C | 7363 hCoV-19/USA/ID-UW-4597/2020\|EPI_ISL_460628\|2020-03 | ···················· | ···················· | ···················· | ···················· | ···················· | ···················· | |
| C | 7364 hCoV-19/USA/ID-UW-4600/2020\|EPI_ISL_460629\|2020-03 | ···················· | ···················· | ···················· | ···················· | ···················· | ···················· | |
| C | 7365 hCoV-19/USA/WA-UW-4619/2020\|EPI_ISL_460630\|2020-03 | ···················· | ···················· | ···················· | ···················· | ···················· | ···················· | |
| C | 7366 hCoV-19/USA/WA-UW-4633/2020\|EPI_ISL_460632\|2020-03 | ···················· | ···················· | ···················· | ···················· | ···················· | ···················· | |
| C | 7367 hCoV-19/USA/WA-UW-4643/2020\|EPI_ISL_460633\|2020-04 | ···················· | ···················· | ···················· | ···················· | ···················· | ···················· | |
| C | 7368 hCoV-19/England/WARW04/2020\|EPI_ISL_460635\|2020-04 | ···················· | ···················· | ···················· | ···················· | ···················· | ···················· | |
| C | 7369 hCoV-19/Netherlands/Drenthe_2/2020\|EPI_ISL_460636\| | ···················· | ···················· | ···················· | ···················· | ···················· | ···················· | |
| C | 7370 hCoV-19/Netherlands/Drenthe_3/2020\|EPI_ISL_460637\| | ···················· | ···················· | ···················· | ···················· | ···················· | ···················· | |
| C | 7371 hCoV-19/Netherlands/Flevoland_5/2020\|EPI_ISL_46063 | ···················· | ···················· | ···················· | ···················· | ···················· | ···················· | |
| C | 7372 hCoV-19/Netherlands/Flevoland_6/2020\|EPI_ISL_46063 | ···················· | ···················· | ···················· | ···················· | ···················· | ···················· | |
| C | 7373 hCoV-19/Netherlands/Flevoland_7/2020\|EPI_ISL_46064 | ···················· | ···················· | ···················· | ···················· | ···················· | ···················· | |
| C | 7374 hCoV-19/Netherlands/Friesland_19/2020\|EPI_ISL_4606 | ···················· | ···················· | ···················· | ···················· | ···················· | ···················· | |
| C | 7375 hCoV-19/Netherlands/Friesland_20/2020\|EPI_ISL_4606 | ···················· | ···················· | ···················· | ···················· | ···················· | ···················· | |
| C | 7376 hCoV-19/Netherlands/Friesland_21/2020\|EPI_ISL_4606 | ···················· | ···················· | ···················· | ···················· | ···················· | ···················· | |
| C | 7377 hCoV-19/Netherlands/Friesland_22/2020\|EPI_ISL_4606 | ···················· | ···················· | ···················· | ···················· | ···················· | ···················· | |
| C | 7378 hCoV-19/Netherlands/Friesland_24/2020\|EPI_ISL_4606 | ···················· | ···················· | ···················· | ···················· | ···················· | ···················· | |
| C | 7379 hCoV-19/Netherlands/Friesland_25/2020\|EPI_ISL_4606 | ···················· | ···················· | ···················· | ···················· | ···················· | ···················· | |
| C | 7380 hCoV-19/Netherlands/Friesland_26/2020\|EPI_ISL_4606 | ···················· | ···················· | ···················· | ···················· | ···················· | ···················· | |
| C | 7381 hCoV-19/Netherlands/Friesland_27/2020\|EPI_ISL_4606 | ···················· | ···················· | ···················· | ···················· | ···················· | ···················· | |
| C | 7382 hCoV-19/England/NOTT-111618/2020\|EPI_ISL_461900\|20 | ···················· | ···················· | ···················· | ···················· | ···················· | ···················· | |
| C | 7383 hCoV-19/England/NOTT-111627/2020\|EPI_ISL_461901\|20 | ···················· | ···················· | ···················· | ···················· | ···················· | ···················· | |
| C | 7384 hCoV-19/England/NOTT-111636/2020\|EPI_ISL_461902\|20 | ···················· | ···················· | ···················· | ···················· | ···················· | ···················· | |
| C | 7385 hCoV-19/England/NOTT-111681/2020\|EPI_ISL_461905\|20 | ···················· | ···················· | ···················· | ···················· | ···················· | ···················· | |
| C | 7386 hCoV-19/England/NOTT-111690/2020\|EPI_ISL_461906\|20 | ···················· | ···················· | ···················· | ···················· | ···················· | ···················· | |
| C | 7387 hCoV-19/England/NOTT-1116AF/2020\|EPI_ISL_461907\|20 | ···················· | ···················· | ···················· | ···················· | ···················· | ···················· | |
| C | 7388 hCoV-19/England/NOTT-1116CD/2020\|EPI_ISL_461908\|20 | ···················· | ···················· | ···················· | ···················· | ···················· | ···················· | |
| C | 7389 hCoV-19/England/NOTT-1116DC/2020\|EPI_ISL_461909\|20 | ···················· | ···················· | ···················· | ···················· | ···················· | ···················· | |
| C | 7390 hCoV-19/England/NOTT-1116FA/2020\|EPI_ISL_461911\|20 | ···················· | ···················· | ···················· | ···················· | ···················· | ···················· | |
| C | 7391 hCoV-19/England/NOTT-111706/2020\|EPI_ISL_461912\|20 | ···················· | ···················· | ···················· | ···················· | ···················· | ···················· | |
| C | 7392 hCoV-19/England/NOTT-111733/2020\|EPI_ISL_461915\|20 | ···················· | ···················· | ···················· | ···················· | ···················· | ···················· | |
| C | 7393 hCoV-19/England/NOTT-111742/2020\|EPI_ISL_461916\|20 | ···················· | ···················· | ···················· | ···················· | ···················· | ···················· | |
| C | 7394 hCoV-19/England/NOTT-111900/2020\|EPI_ISL_461940\|20 | ···················· | ···················· | ···················· | ···················· | ···················· | ···················· | |
| C | 7395 hCoV-19/England/NOTT-11192E/2020\|EPI_ISL_461942\|20 | ···················· | ···················· | ···················· | ···················· | ···················· | ···················· | |
| C | 7396 hCoV-19/England/NOTT-11193D/2020\|EPI_ISL_461943\|20 | ···················· | ···················· | ···················· | ···················· | ···················· | ···················· | |
| C | 7397 hCoV-19/England/NOTT-111979/2020\|EPI_ISL_461945\|20 | ···················· | ···················· | ···················· | ···················· | ···················· | ···················· | |
| C | 7398 hCoV-19/England/NOTT-111997/2020\|EPI_ISL_461946\|20 | ···················· | ···················· | ···················· | ···················· | ···················· | ···················· | |
| C | 7399 hCoV-19/England/NOTT-1119A6/2020\|EPI_ISL_461947\|20 | ···················· | ···················· | ···················· | ···················· | ···················· | ···················· | |
| C | 7400 hCoV-19/England/NOTT-1119B5/2020\|EPI_ISL_461948\|20 | ···················· | ···················· | ···················· | ···················· | ···················· | ···················· | |
| C | 7401 hCoV-19/England/NOTT-1119C4/2020\|EPI_ISL_461949\|20 | ···················· | ···················· | ···················· | ···················· | ···················· | ···················· | |
| C | 7402 hCoV-19/England/NOTT-1119F1/2020\|EPI_ISL_461950\|20 | ···················· | ···················· | ···················· | ···················· | ···················· | ···················· | |
| C | 7403 hCoV-19/England/NOTT-111A0D/2020\|EPI_ISL_461951\|20 | ···················· | ···················· | ···················· | ···················· | ···················· | ···················· | |
| C | 7404 hCoV-19/England/NOTT-111A2B/2020\|EPI_ISL_461952\|20 | ···················· | ···················· | ···················· | ···················· | ···················· | ···················· | |

| | | | | | | | | | |
|---|---|---|---|---|---|---|---|---|---|
| C | 7405 | hCoV-19/England/NOTT-111A3A/2020\|EPI_ISL_461953\|20 | .................... | .................... | .................... | .................... | .................... | .................... | |
| C | 7406 | hCoV-19/England/NOTT-111A49/2020\|EPI_ISL_461954\|20 | .................... | .................... | .................... | .................... | .................... | .................... | |
| C | 7407 | hCoV-19/England/NOTT-111AB2/2020\|EPI_ISL_461958\|20 | .................... | .................... | .................... | .................... | .................... | .................... | |
| C | 7408 | hCoV-19/England/NOTT-111AEF/2020\|EPI_ISL_461960\|20 | .................... | .................... | .................... | .................... | .................... | .................... | |
| C | 7409 | hCoV-19/England/NOTT-111B0A/2020\|EPI_ISL_461961\|20 | .................... | .................... | .................... | .................... | .................... | .................... | |
| C | 7410 | hCoV-19/England/NOTT-111B28/2020\|EPI_ISL_461962\|20 | .................... | .................... | .................... | .................... | .................... | .................... | |
| C | 7411 | hCoV-19/England/NOTT-111B37/2020\|EPI_ISL_461963\|20 | .................... | .................... | .................... | .................... | .................... | .................... | |
| C | 7412 | hCoV-19/England/NOTT-111B46/2020\|EPI_ISL_461964\|20 | .................... | .................... | .................... | .................... | .................... | .................... | |
| C | 7413 | hCoV-19/England/NOTT-111B64/2020\|EPI_ISL_461965\|20 | .................... | .................... | .................... | .................... | .................... | .................... | |
| C | 7414 | hCoV-19/England/NOTT-111B82/2020\|EPI_ISL_461967\|20 | .................... | .................... | .................... | .................... | .................... | .................... | |
| C | 7415 | hCoV-19/England/NOTT-111BA0/2020\|EPI_ISL_461968\|20 | .................... | .................... | .................... | .................... | .................... | .................... | |
| C | 7416 | hCoV-19/England/NOTT-111BBF/2020\|EPI_ISL_461969\|20 | .................... | .................... | .................... | .................... | .................... | .................... | |
| C | 7417 | hCoV-19/England/PORT-2CF645/2020\|EPI_ISL_461971\|20 | .................... | .................... | .................... | .................... | .................... | .................... | |
| C | 7418 | hCoV-19/England/PORT-2CF6AF/2020\|EPI_ISL_461972\|20 | .................... | .................... | .................... | .................... | .................... | .................... | |
| C | 7419 | hCoV-19/England/PORT-2CF6BE/2020\|EPI_ISL_461973\|20 | .................... | .................... | .................... | .................... | .................... | .................... | |
| C | 7420 | hCoV-19/England/PORT-2CF6CD/2020\|EPI_ISL_461974\|20 | .................... | .................... | .................... | .................... | .................... | .................... | |
| C | 7421 | hCoV-19/England/PORT-2CF6DC/2020\|EPI_ISL_461975\|20 | .................... | .................... | .................... | .................... | .................... | .................... | |
| C | 7422 | hCoV-19/England/PORT-2CF6EB/2020\|EPI_ISL_461976\|20 | .................... | .................... | .................... | .................... | .................... | .................... | |
| C | 7423 | hCoV-19/England/PORT-2CF715/2020\|EPI_ISL_461977\|20 | .................... | .................... | .................... | .................... | .................... | .................... | |
| C | 7424 | hCoV-19/England/PORT-2CF742/2020\|EPI_ISL_461979\|20 | .................... | .................... | .................... | .................... | .................... | .................... | |
| C | 7425 | hCoV-19/England/NORW-E811A/2020\|EPI_ISL_449110\|202 | .................... | .................... | .................... | .................... | .................... | .................... | |
| C | 7426 | hCoV-19/England/NORW-E8129/2020\|EPI_ISL_449111\|202 | .................... | .................... | .................... | .................... | .................... | .................... | |
| C | 7427 | hCoV-19/England/NORW-E8138/2020\|EPI_ISL_449112\|202 | .................... | .................... | .................... | .................... | .................... | .................... | |
| C | 7428 | hCoV-19/England/NORW-E8165/2020\|EPI_ISL_449115\|202 | .................... | .................... | .................... | .................... | .................... | .................... | |
| C | 7429 | hCoV-19/England/NORW-E8174/2020\|EPI_ISL_449116\|202 | .................... | .................... | .................... | .................... | .................... | .................... | |
| C | 7430 | hCoV-19/England/NORW-E8183/2020\|EPI_ISL_449117\|202 | .................... | .................... | .................... | .................... | .................... | .................... | |
| C | 7431 | hCoV-19/England/NORW-E81A1/2020\|EPI_ISL_449118\|202 | .................... | .................... | .................... | .................... | .................... | .................... | |
| C | 7432 | hCoV-19/England/NORW-E81B0/2020\|EPI_ISL_449119\|202 | .................... | .................... | .................... | .................... | .................... | .................... | |
| C | 7433 | hCoV-19/England/NORW-E81DE/2020\|EPI_ISL_449121\|202 | .................... | .................... | .................... | .................... | .................... | .................... | |
| C | 7434 | hCoV-19/England/NORW-E81ED/2020\|EPI_ISL_449122\|202 | .................... | .................... | .................... | .................... | .................... | .................... | |
| C | 7435 | hCoV-19/England/NORW-E8208/2020\|EPI_ISL_449124\|202 | .................... | .................... | .................... | .................... | .................... | .................... | |
| C | 7436 | hCoV-19/England/NORW-E8217/2020\|EPI_ISL_449125\|202 | .................... | .................... | .................... | .................... | .................... | .................... | |
| C | 7437 | hCoV-19/England/NORW-E8226/2020\|EPI_ISL_449126\|202 | .................... | .................... | .................... | .................... | .................... | .................... | |
| C | 7438 | hCoV-19/England/NORW-E8235/2020\|EPI_ISL_449127\|202 | .................... | .................... | .................... | .................... | .................... | .................... | |
| C | 7439 | hCoV-19/England/NORW-E8244/2020\|EPI_ISL_449128\|202 | .................... | .................... | .................... | .................... | .................... | .................... | |
| C | 7440 | hCoV-19/England/NORW-E8253/2020\|EPI_ISL_449129\|202 | .................... | .................... | .................... | .................... | .................... | .................... | |
| C | 7441 | hCoV-19/England/NORW-E8262/2020\|EPI_ISL_449130\|202 | .................... | .................... | .................... | .................... | .................... | .................... | |
| C | 7442 | hCoV-19/England/NORW-E8271/2020\|EPI_ISL_449131\|202 | .................... | .................... | .................... | .................... | .................... | .................... | |
| C | 7443 | hCoV-19/England/NORW-E8280/2020\|EPI_ISL_449132\|202 | .................... | .................... | .................... | .................... | .................... | .................... | |
| C | 7444 | hCoV-19/England/NORW-E829F/2020\|EPI_ISL_449133\|202 | .................... | .................... | .................... | .................... | .................... | .................... | |
| C | 7445 | hCoV-19/England/NORW-E82BD/2020\|EPI_ISL_449135\|202 | .................... | .................... | .................... | .................... | .................... | .................... | |
| C | 7446 | hCoV-19/England/NORW-E82CC/2020\|EPI_ISL_449136\|202 | .................... | .................... | .................... | .................... | .................... | .................... | |
| C | 7447 | hCoV-19/England/NORW-E82DB/2020\|EPI_ISL_449137\|202 | .................... | .................... | .................... | .................... | .................... | .................... | |
| C | 7448 | hCoV-19/England/NORW-E82EA/2020\|EPI_ISL_449138\|202 | .................... | .................... | .................... | .................... | .................... | .................... | |
| C | 7449 | hCoV-19/England/NORW-E82F9/2020\|EPI_ISL_449139\|202 | .................... | .................... | .................... | .................... | .................... | .................... | |
| C | 7450 | hCoV-19/England/NORW-E8305/2020\|EPI_ISL_449140\|202 | .................... | .................... | .................... | .................... | .................... | .................... | |
| C | 7451 | hCoV-19/England/NORW-E8314/2020\|EPI_ISL_449141\|202 | .................... | .................... | .................... | .................... | .................... | .................... | |
| C | 7452 | hCoV-19/England/NORW-E8332/2020\|EPI_ISL_449142\|202 | .................... | .................... | .................... | .................... | .................... | .................... | |
| C | 7453 | hCoV-19/England/NORW-E838D/2020\|EPI_ISL_449143\|202 | .................... | .................... | .................... | .................... | .................... | .................... | |
| C | 7454 | hCoV-19/England/NORW-E839C/2020\|EPI_ISL_449144\|202 | .................... | .................... | .................... | .................... | .................... | .................... | |
| C | 7455 | hCoV-19/England/NORW-E83AB/2020\|EPI_ISL_449145\|202 | .................... | .................... | .................... | .................... | .................... | .................... | |
| C | 7456 | hCoV-19/England/NORW-E83C9/2020\|EPI_ISL_449146\|202 | .................... | .................... | .................... | .................... | .................... | .................... | |
| C | 7457 | hCoV-19/England/NORW-E83F6/2020\|EPI_ISL_449148\|202 | .................... | .................... | .................... | .................... | .................... | .................... | |
| C | 7458 | hCoV-19/England/NORW-E8402/2020\|EPI_ISL_449149\|202 | .................... | .................... | .................... | .................... | .NN................. | .................... | |
| C | 7459 | hCoV-19/England/NORW-E8411/2020\|EPI_ISL_449150\|202 | .................... | .................... | .................... | .................... | .................... | .................... | |
| C | 7460 | hCoV-19/England/NORW-E8420/2020\|EPI_ISL_449151\|202 | .................... | .................... | .................... | .................... | .................... | .................... | |
| C | 7461 | hCoV-19/England/NORW-E844E/2020\|EPI_ISL_449152\|202 | .................... | .................... | .................... | .................... | .................... | .................... | |
| C | 7462 | hCoV-19/England/NORW-E8499/2020\|EPI_ISL_449153\|202 | .................... | .................... | .................... | .................... | .................... | .................... | |
| C | 7463 | hCoV-19/England/NORW-E84A8/2020\|EPI_ISL_449154\|202 | .................... | .................... | .................... | .................... | .................... | .................... | |
| C | 7464 | hCoV-19/England/NORW-E84B7/2020\|EPI_ISL_449155\|202 | .................... | .................... | .................... | .................... | .................... | .................... | |
| C | 7465 | hCoV-19/England/NORW-E84D5/2020\|EPI_ISL_449156\|202 | .................... | .................... | .................... | .................... | .................... | .................... | |
| C | 7466 | hCoV-19/England/NORW-E850F/2020\|EPI_ISL_449157\|202 | .................... | .................... | .................... | .................... | .................... | .................... | |
| C | 7467 | hCoV-19/England/NORW-E851E/2020\|EPI_ISL_449158\|202 | .................... | .................... | .................... | .................... | .................... | .................... | |
| C | 7468 | hCoV-19/England/NORW-E852D/2020\|EPI_ISL_449159\|202 | .................... | .................... | .................... | .................... | .................... | .................... | |
| C | 7469 | hCoV-19/England/NORW-E85C3/2020\|EPI_ISL_449164\|202 | .................... | .................... | .................... | .................... | .................... | .................... | |
| C | 7470 | hCoV-19/England/NORW-E85E1/2020\|EPI_ISL_449165\|202 | .................... | .................... | .................... | .................... | .................... | .................... | |
| C | 7471 | hCoV-19/England/NORW-E862A/2020\|EPI_ISL_449167\|202 | .................... | .................... | .................... | .................... | ..NN................ | .................... | |
| C | 7472 | hCoV-19/England/NORW-E8639/2020\|EPI_ISL_449168\|202 | .................... | .................... | .................... | .................... | .................... | .................... | |
| C | 7473 | hCoV-19/England/NORW-E8648/2020\|EPI_ISL_449169\|202 | .................... | .................... | .................... | .................... | .................... | .................... | |
| C | 7474 | hCoV-19/England/NORW-E8666/2020\|EPI_ISL_449170\|202 | .................... | .................... | .................... | .................... | .................... | .................... | |
| C | 7475 | hCoV-19/England/NORW-E8675/2020\|EPI_ISL_449171\|202 | .................... | .................... | .................... | .................... | .................... | .................... | |
| C | 7476 | hCoV-19/England/NORW-E8684/2020\|EPI_ISL_449172\|202 | .................... | .................... | .................... | .................... | .................... | .................... | |
| C | 7477 | hCoV-19/England/NORW-E8693/2020\|EPI_ISL_449173\|202 | .................... | .................... | .................... | .................... | .................... | .................... | |
| C | 7478 | hCoV-19/England/NORW-E86A2/2020\|EPI_ISL_449174\|202 | .................... | .................... | .................... | .................... | .................... | .................... | |
| C | 7480 | hCoV-19/Scotland/CVR2983/2020\|EPI_ISL_449176\|2020- | .................... | ...................A | .................... | .................... | .................... | .................... | 1 |
| C | 7481 | hCoV-19/Scotland/CVR2985/2020\|EPI_ISL_449178\|2020- | .................... | ...................A | .................... | .................... | .................... | .................... | 1 |
| C | 7482 | hCoV-19/Scotland/CVR2986/2020\|EPI_ISL_449179\|2020- | .................... | .................... | .................... | .................... | .................... | .................... | |
| C | 7483 | hCoV-19/Scotland/CVR3142/2020\|EPI_ISL_449187\|2020- | .................... | ..............C..... | .................... | .................... | .................... | .................... | |

| | | | | | | | | |
|---|---|---|---|---|---|---|---|---|
| C | 7484 hCoV-19/Scotland/CVR3144/2020\|EPI_ISL_449188\|2020- | ···················· | ···················A | ···················· | ···················· | ···················· | ···················· | 1 |
| C | 7485 hCoV-19/Scotland/CVR3146/2020\|EPI_ISL_449189\|2020- | ···················· | ···················· | ···················· | ···················· | ···················· | ···················· | |
| C | 7486 hCoV-19/USA/NY-PV09421/2020\|EPI_ISL_450100\|2020-03 | ···················· | ···················· | ···················· | ···················· | ···················· | ···················· | |
| C | 7487 hCoV-19/USA/NY-PV09422/2020\|EPI_ISL_450101\|2020-03 | ···················· | ···················· | ···················· | ···················· | ···················· | ···················· | |
| C | 7488 hCoV-19/USA/NY-PV09423/2020\|EPI_ISL_450102\|2020-03 | ···················· | ···················· | ···················· | ···················· | ···················· | ···················· | |
| C | 7489 hCoV-19/USA/NY-PV09425/2020\|EPI_ISL_450103\|2020-03 | ···················· | ···················· | ···················· | ···················· | ···················· | ···················· | |
| C | 7490 hCoV-19/USA/NY-PV09427/2020\|EPI_ISL_450104\|2020-03 | ···················· | ···················· | ···················· | ···················· | ···················· | ···················· | |
| C | 7491 hCoV-19/USA/NY-PV09428/2020\|EPI_ISL_450105\|2020-03 | ···················· | ···················· | ···················· | ···················· | ···················· | ···················· | |
| C | 7492 hCoV-19/USA/NY-PV09429/2020\|EPI_ISL_450106\|2020-03 | ···················· | ···················· | ···················· | ···················· | ···················· | ···················· | |
| C | 7493 hCoV-19/USA/NY-PV09430/2020\|EPI_ISL_450107\|2020-03 | ···················· | ···················· | ···················· | ···················· | ···················· | ···················· | |
| C | 7494 hCoV-19/USA/NY-PV09431/2020\|EPI_ISL_450108\|2020-03 | ···················· | ···················· | ···················· | ···················· | ···················· | ···················· | |
| C | 7495 hCoV-19/USA/NY-PV09432/2020\|EPI_ISL_450109\|2020-03 | ···················· | ···················· | ···················· | ···················· | ···················· | ···················· | |
| C | 7496 hCoV-19/USA/NY-PV09433/2020\|EPI_ISL_450110\|2020-03 | ···················· | ···················· | ···················· | ···················· | ···················· | ···················· | |
| C | 7497 hCoV-19/USA/NY-PV09434/2020\|EPI_ISL_450111\|2020-03 | ···················· | ···················· | ···················· | ···················· | ···················· | ···················· | |
| C | 7498 hCoV-19/USA/NY-PV09435/2020\|EPI_ISL_450112\|2020-03 | ···················· | ···················· | ···················· | ···················· | ···················· | ···················· | |
| C | 7499 hCoV-19/USA/NY-PV09436/2020\|EPI_ISL_450113\|2020-03 | ···················· | ···················· | ···················· | ···················· | ···················· | ···················· | |
| C | 7500 hCoV-19/USA/NY-PV09437/2020\|EPI_ISL_450114\|2020-03 | ···················· | ···················· | ···················· | ···················· | ···················· | ···················· | |
| C | 7501 hCoV-19/USA/NY-PV09438/2020\|EPI_ISL_450115\|2020-03 | ···················· | ···················· | ···················· | ···················· | ···················· | ···················· | |
| C | 7502 hCoV-19/USA/NY-PV09439/2020\|EPI_ISL_450116\|2020-03 | ···················· | ···················· | ···················· | ···················· | ···················· | ···················· | |
| C | 7503 hCoV-19/USA/NY-PV09440/2020\|EPI_ISL_450117\|2020-03 | ···················· | ···················· | ···················· | ···················· | ···················· | ···················· | |
| C | 7504 hCoV-19/USA/NY-PV09441/2020\|EPI_ISL_450118\|2020-03 | ···················· | ···················· | ···················· | ···················· | ···················· | ···················· | |
| C | 7505 hCoV-19/USA/NY-PV09444/2020\|EPI_ISL_450120\|2020-03 | ···················· | ···················· | ···················· | ···················· | ···················· | ···················· | |
| C | 7506 hCoV-19/USA/NY-PV09446/2020\|EPI_ISL_450121\|2020-03 | ···················· | ···················· | ···················· | ···················· | ···················· | ···················· | |
| C | 7507 hCoV-19/USA/NY-PV09449/2020\|EPI_ISL_450123\|2020-03 | ···················· | ···················· | ···················· | ···················· | ···················· | ···················· | |
| C | 7508 hCoV-19/USA/NY-PV09450/2020\|EPI_ISL_450124\|2020-03 | ···················· | ···················· | ···················· | ···················· | ···················· | ···················· | |
| C | 7509 hCoV-19/USA/NY-PV09451/2020\|EPI_ISL_450125\|2020-03 | ···················· | ···················· | ···················· | ···················· | ···················· | ···················· | |
| C | 7510 hCoV-19/USA/NY-PV09452/2020\|EPI_ISL_450126\|2020-03 | ···················· | ···················· | ···················· | ···················· | ···················· | ···················· | |
| C | 7511 hCoV-19/USA/NY-PV09453/2020\|EPI_ISL_450127\|2020-03 | ···················· | ···················· | ···················· | ···················· | ···················· | ···················· | |
| C | 7512 hCoV-19/USA/NY-PV09454/2020\|EPI_ISL_450128\|2020-03 | ···················· | ···················· | ···················· | ···················· | ···················· | ···················· | |
| C | 7513 hCoV-19/USA/NY-PV09455/2020\|EPI_ISL_450129\|2020-03 | ···················· | ···················· | ···················· | ···················· | ···················· | ···················· | |
| C | 7514 hCoV-19/USA/NY-PV09456/2020\|EPI_ISL_450130\|2020-03 | ···················· | ···················· | ···················· | ···················· | ···················· | ···················· | |
| C | 7515 hCoV-19/USA/NY-PV09459/2020\|EPI_ISL_450132\|2020-03 | ···················· | ···················· | ···················· | ···················· | ···················· | ···················· | |
| C | 7516 hCoV-19/USA/NY-PV09460/2020\|EPI_ISL_450133\|2020-03 | ···················· | ···················· | ···················· | ···················· | ···················· | ···················· | |
| C | 7517 hCoV-19/USA/NY-PV09461/2020\|EPI_ISL_450134\|2020-03 | ···················· | ···················· | ···················· | ···················· | ···················· | ···················· | |
| C | 7518 hCoV-19/USA/NY-PV09462/2020\|EPI_ISL_450135\|2020-03 | ···················· | ···················· | ···················· | ···················· | ···················· | ···················· | |
| C | 7519 hCoV-19/USA/NY-PV09463/2020\|EPI_ISL_450136\|2020-03 | ···················· | ···················· | ···················· | ···················· | ···················· | ···················· | |
| C | 7520 hCoV-19/USA/NY-PV09464/2020\|EPI_ISL_450137\|2020-03 | ···················· | ···················· | ···················· | ···················· | ···················· | ···················· | |
| C | 7521 hCoV-19/USA/NJ-PV09465/2020\|EPI_ISL_450138\|2020-03 | ···················· | ···················· | ···················· | ···················· | ···················· | ···················· | |
| C | 7522 hCoV-19/USA/NY-PV09466/2020\|EPI_ISL_450139\|2020-03 | ···················· | ···················· | ···················· | ···················· | ···················· | ···················· | |
| C | 7523 hCoV-19/USA/NY-PV09472/2020\|EPI_ISL_450144\|2020-03 | ···················· | ···················· | ···················· | ···················· | ···················· | ···················· | |
| C | 7524 hCoV-19/USA/NJ-PV09473/2020\|EPI_ISL_450145\|2020-03 | ···················· | ···················· | ···················· | ···················· | ···················· | ···················· | |
| C | 7525 hCoV-19/USA/NY-PV09474/2020\|EPI_ISL_450146\|2020-03 | ···················· | ···················· | ···················· | ···················· | ···················· | ···················· | |
| C | 7526 hCoV-19/USA/NY-PV09475/2020\|EPI_ISL_450147\|2020-03 | ···················· | ···················· | ···················· | ···················· | ···················· | ···················· | |
| C | 7527 hCoV-19/USA/NY-PV09476/2020\|EPI_ISL_450148\|2020-03 | ···················· | ···················· | ···················· | ···················· | ···················· | ···················· | |
| C | 7528 hCoV-19/USA/NY-PV09477/2020\|EPI_ISL_450149\|2020-03 | ···················· | ···················· | ···················· | ···················· | ···················· | ···················· | |
| C | 7529 hCoV-19/USA/NY-PV09478/2020\|EPI_ISL_450150\|2020-03 | ···················· | ···················· | ···················· | ···················· | ···················· | ···················· | |
| C | 7530 hCoV-19/USA/NY-PV09479/2020\|EPI_ISL_450151\|2020-03 | ···················· | ···················· | ···················· | ···················· | ···················· | ···················· | |
| C | 7531 hCoV-19/USA/NY-PV09480/2020\|EPI_ISL_450152\|2020-03 | ···················· | ···················· | ···················· | ···················· | ···················· | ···················· | |
| C | 7532 hCoV-19/USA/NY-PV09481/2020\|EPI_ISL_450153\|2020-03 | ···················· | ···················· | ···················· | ···················· | ···················· | ···················· | |
| C | 7533 hCoV-19/USA/NY-PV09482/2020\|EPI_ISL_450154\|2020-03 | ···················· | ···················· | ···················· | ···················· | ···················· | ···················· | |
| C | 7534 hCoV-19/USA/NY-PV09483/2020\|EPI_ISL_450155\|2020-03 | ···················· | ···················· | ···················· | ···················· | ···················· | ···················· | |
| C | 7535 hCoV-19/USA/NY-PV09484/2020\|EPI_ISL_450156\|2020-03 | ···················· | ···················· | ···················· | ···················· | ···················· | ···················· | |
| C | 7536 hCoV-19/USA/NY-PV09485/2020\|EPI_ISL_450157\|2020-03 | ···················· | ···················· | ···················· | ···················· | ···················· | ···················· | |
| C | 7537 hCoV-19/USA/NY-PV09486/2020\|EPI_ISL_450158\|2020-03 | ···················· | ···················· | ···················· | ···················· | ···················· | ···················· | |
| C | 7538 hCoV-19/USA/NY-PV09487/2020\|EPI_ISL_450159\|2020-03 | ···················· | ···················· | ···················· | ···················· | ···················· | ···················· | |
| C | 7539 hCoV-19/USA/NY-PV09488/2020\|EPI_ISL_450160\|2020-03 | ···················· | ···················· | ···················· | ···················· | ···················· | ···················· | |
| C | 7540 hCoV-19/USA/NY-PV09489/2020\|EPI_ISL_450161\|2020-03 | ···················· | ···················· | ···················· | ···················· | ···················· | ···················· | |
| C | 7541 hCoV-19/USA/NY-PV09490/2020\|EPI_ISL_450162\|2020-03 | ···················· | ···················· | ···················· | ···················· | ···················· | ···················· | |
| C | 7542 hCoV-19/USA/NY-PV09491/2020\|EPI_ISL_450163\|2020-03 | ···················· | ···················· | ···················· | ···················· | ···················· | ···················· | |
| C | 7543 hCoV-19/USA/NY-PV09492/2020\|EPI_ISL_450164\|2020-03 | ···················· | ···················· | ···················· | ···················· | ···················· | ···················· | |
| C | 7544 hCoV-19/USA/NY-PV09493/2020\|EPI_ISL_450165\|2020-03 | ···················· | ···················· | ···················· | ···················· | ···················· | ···················· | |
| C | 7545 hCoV-19/USA/NY-PV09494/2020\|EPI_ISL_450166\|2020-03 | ···················· | ···················· | ···················· | ···················· | ···················· | ···················· | |
| C | 7546 hCoV-19/USA/NY-PV09495/2020\|EPI_ISL_450167\|2020-03 | ···················· | ···················· | ···················· | ···················· | ···················· | ···················· | |
| C | 7547 hCoV-19/USA/NY-PV09496/2020\|EPI_ISL_450168\|2020-03 | ···················· | ···················· | ···················· | ···················· | ···················· | ···················· | |
| C | 7548 hCoV-19/USA/NY-PV09497/2020\|EPI_ISL_450169\|2020-03 | ···················· | ···················· | ···················· | ···················· | ···················· | ···················· | |
| C | 7549 hCoV-19/USA/NY-PV09504/2020\|EPI_ISL_450170\|2020-03 | ···················· | ······A············· | ···················· | ···················· | ···················· | ···················· | |
| C | 7550 hCoV-19/USA/LA-SR0318/2020\|EPI_ISL_450173\|2020-04- | ···················· | ···················· | ···················· | ···················· | ···················· | ···················· | |
| C | 7551 hCoV-19/USA/LA-SR0319/2020\|EPI_ISL_450174\|2020-04- | ···················· | ···················· | ···················· | ···················· | ···················· | ···················· | |
| C | 7552 hCoV-19/USA/LA-SR0320/2020\|EPI_ISL_450175\|2020-04- | ···················· | ···················· | ···················· | ···················· | ···················· | ···················· | |
| C | 7553 hCoV-19/USA/LA-SR0322/2020\|EPI_ISL_450176\|2020-04- | ···················· | ···················· | ···················· | ···················· | ···················· | ···················· | |
| C | 7554 hCoV-19/USA/NY-NYUMC705/2020\|EPI_ISL_451420\|2020-0 | ···················· | ···················· | ···················· | ···················· | ···················· | ···················· | |
| C | 7555 hCoV-19/USA/NY-NYUMC706/2020\|EPI_ISL_451421\|2020-0 | ···················· | ···················· | ···················· | ···················· | ···················· | ···················· | |
| C | 7556 hCoV-19/USA/NY-NYUMC707/2020\|EPI_ISL_451422\|2020-0 | ···················· | ···················· | ···················· | ···················· | ···················· | ···················· | |
| C | 7557 hCoV-19/USA/NY-NYUMC708/2020\|EPI_ISL_451423\|2020-0 | ···················· | ···················· | ···················· | ···················· | ···················· | ···················· | |
| C | 7558 hCoV-19/USA/NY-NYUMC709/2020\|EPI_ISL_451424\|2020-0 | ···················· | ···················· | ···················· | ···················· | ···················· | ···················· | |
| C | 7559 hCoV-19/USA/NY-NYUMC711/2020\|EPI_ISL_451426\|2020-0 | ···················· | ···················· | ···················· | ···················· | ···················· | ···················· | |
| C | 7560 hCoV-19/USA/NY-NYUMC712/2020\|EPI_ISL_451427\|2020-0 | ···················· | ···················· | ···················· | ···················· | ···················· | ···················· | |
| C | 7561 hCoV-19/USA/NY-NYUMC713/2020\|EPI_ISL_451428\|2020-0 | ···················· | ···················· | ···················· | ···················· | ···················· | ···················· | |

| | | | | | | | | |
|---|---|---|---|---|---|---|---|---|
| C | 7562 | hCoV-19/USA/NY-NYUMC714/2020\|EPI_ISL_451429\|2020-0 | .................... | .................... | ..................... | .................... | .................... | .................... |
| C | 7563 | hCoV-19/USA/NY-NYUMC715/2020\|EPI_ISL_451430\|2020-0 | .................... | .................... | ..................... | .................... | .................... | .................... |
| C | 7564 | hCoV-19/USA/NY-NYUMC716/2020\|EPI_ISL_451431\|2020-0 | .................... | .................... | ..................... | .................... | .................... | .................... |
| C | 7565 | hCoV-19/USA/NY-NYUMC717/2020\|EPI_ISL_451432\|2020-0 | .................... | .................... | ..................... | .................... | .................... | .................... |
| C | 7566 | hCoV-19/USA/NY-NYUMC719/2020\|EPI_ISL_451434\|2020-0 | .................... | .................... | ..................... | .................... | .................... | .................... |
| C | 7567 | hCoV-19/USA/NY-NYUMC720/2020\|EPI_ISL_451435\|2020-0 | .................... | .................... | ..................... | .................... | .................... | .................... |
| C | 7568 | hCoV-19/USA/NY-NYUMC721/2020\|EPI_ISL_451436\|2020-0 | .................... | .................... | ..................... | .................... | .................... | .................... |
| C | 7569 | hCoV-19/USA/NY-NYUMC722/2020\|EPI_ISL_451437\|2020-0 | .................... | .................... | ..................... | .................... | .................... | .................... |
| C | 7570 | hCoV-19/USA/NY-NYUMC723/2020\|EPI_ISL_451438\|2020-0 | .................... | .................... | ..................... | .................... | .................... | .................... |
| C | 7571 | hCoV-19/USA/NY-NYUMC724/2020\|EPI_ISL_451439\|2020-0 | .................... | .................... | ..................... | .................... | .................... | .................... |
| C | 7572 | hCoV-19/USA/NY-NYUMC725/2020\|EPI_ISL_451440\|2020-0 | .................... | .................... | ..................... | .................... | .................... | .................... |
| C | 7573 | hCoV-19/USA/NY-NYUMC726/2020\|EPI_ISL_451441\|2020-0 | .................... | .................... | ..................... | .................... | .................... | .................... |
| C | 7574 | hCoV-19/USA/NY-NYUMC727/2020\|EPI_ISL_451442\|2020-0 | .................... | .................... | ..................... | .................... | .................... | .................... |
| C | 7575 | hCoV-19/USA/NY-NYUMC728/2020\|EPI_ISL_451443\|2020-0 | .................... | .................... | ..................... | .................... | .................... | .................... |
| C | 7576 | hCoV-19/USA/NY-NYUMC729/2020\|EPI_ISL_451444\|2020-0 | .................... | .................... | ..................... | .................... | .................... | .................... |
| C | 7577 | hCoV-19/USA/NY-NYUMC730/2020\|EPI_ISL_451445\|2020-0 | .................... | .................... | ..................... | .................... | .................... | .................... |
| C | 7578 | hCoV-19/USA/NY-NYUMC731/2020\|EPI_ISL_451446\|2020-0 | .................... | .................... | ..................... | .................... | .................... | .................... |
| C | 7579 | hCoV-19/USA/NY-NYUMC732/2020\|EPI_ISL_451447\|2020-0 | .................... | .................... | ..................... | .................... | .................... | .................... |
| C | 7580 | hCoV-19/USA/NY-NYUMC733/2020\|EPI_ISL_451448\|2020-0 | .................... | .................... | ..................... | .................... | .................... | .................... |
| C | 7581 | hCoV-19/USA/NY-NYUMC734/2020\|EPI_ISL_451449\|2020-0 | .................... | .................... | ..................... | .................... | .................... | .................... |
| C | 7582 | hCoV-19/USA/NY-NYUMC735/2020\|EPI_ISL_451450\|2020-0 | .................... | .................... | ..................... | .................... | .................... | .................... |
| C | 7583 | hCoV-19/USA/NY-NYUMC736/2020\|EPI_ISL_451451\|2020-0 | .................... | .................... | ..................... | .................... | .................... | .................... |
| C | 7584 | hCoV-19/USA/NY-NYUMC737/2020\|EPI_ISL_451452\|2020-0 | .................... | .................... | ..................... | .................... | .................... | .................... |
| C | 7585 | hCoV-19/USA/NY-NYUMC738/2020\|EPI_ISL_451453\|2020-0 | .................... | .................... | ..................... | .................... | .................... | .................... |
| C | 7586 | hCoV-19/USA/NY-NYUMC739/2020\|EPI_ISL_451454\|2020-0 | .................... | .................... | ..................... | .................... | .................... | .................... |
| C | 7587 | hCoV-19/USA/NY-NYUMC740/2020\|EPI_ISL_451455\|2020-0 | .................... | .................... | ..................... | .................... | .................... | .................... |
| C | 7588 | hCoV-19/USA/NY-NYUMC741/2020\|EPI_ISL_451456\|2020-0 | .................... | .................... | ..................... | .................... | .................... | .................... |
| C | 7589 | hCoV-19/USA/NY-NYUMC742/2020\|EPI_ISL_451457\|2020-0 | .................... | .................... | ..................... | .................... | .................... | .................... |
| C | 7590 | hCoV-19/USA/NY-NYUMC743/2020\|EPI_ISL_451458\|2020-0 | .................... | .................... | ..................... | .................... | .................... | .................... |
| C | 7591 | hCoV-19/USA/NY-NYUMC744/2020\|EPI_ISL_451459\|2020-0 | .................... | .................... | ..................... | .................... | .................... | .................... |
| C | 7592 | hCoV-19/USA/NY-NYUMC745/2020\|EPI_ISL_451460\|2020-0 | .................... | .................... | ..................... | .................... | .................... | .................... |
| C | 7593 | hCoV-19/USA/NY-NYUMC746/2020\|EPI_ISL_451461\|2020-0 | .................... | .................... | ..................... | .................... | .................... | .................... |
| C | 7594 | hCoV-19/USA/NY-NYUMC747/2020\|EPI_ISL_451462\|2020-0 | .................... | .................... | ..................... | .................... | .................... | .................... |
| C | 7595 | hCoV-19/USA/NY-NYUMC748/2020\|EPI_ISL_451463\|2020-0 | .................... | .................... | ..................... | .................... | .................... | .................... |
| C | 7596 | hCoV-19/USA/NY-NYUMC749/2020\|EPI_ISL_451464\|2020-0 | .................... | .................... | ..................... | .................... | .................... | .................... |
| C | 7597 | hCoV-19/USA/NY-NYUMC750/2020\|EPI_ISL_451465\|2020-0 | .................... | .................... | ..................... | .................... | .................... | .................... |
| C | 7598 | hCoV-19/USA/NY-NYUMC752/2020\|EPI_ISL_451466\|2020-0 | .................... | .................... | ..................... | .................... | .................... | .................... |
| C | 7599 | hCoV-19/USA/NY-NYUMC753/2020\|EPI_ISL_451467\|2020-0 | .................... | .................... | ..................... | .................... | .................... | .................... |
| C | 7600 | hCoV-19/USA/NY-NYUMC761/2020\|EPI_ISL_451475\|2020-0 | .................... | .................... | ..................... | .................... | .................... | .................... |
| C | 7601 | hCoV-19/USA/NY-NYUMC762/2020\|EPI_ISL_451476\|2020-0 | .................... | .................... | ..................... | .................... | .................... | .................... |
| C | 7602 | hCoV-19/USA/NY-NYUMC763/2020\|EPI_ISL_451477\|2020-0 | .................... | .................... | ..................... | .................... | .................... | .................... |
| C | 7603 | hCoV-19/USA/NY-NYUMC764/2020\|EPI_ISL_451478\|2020-0 | .................... | .................... | ..................... | .................... | .................... | .................... |
| C | 7604 | hCoV-19/USA/NY-NYUMC765/2020\|EPI_ISL_451479\|2020-0 | .................... | .................... | ..................... | .................... | .................... | .................... |
| C | 7605 | hCoV-19/Australia/NSW220/2020\|EPI_ISL_451486\|2020- | .................... | .................... | ..................... | .................... | .................... | .................... |
| C | 7606 | hCoV-19/Australia/NSW221/2020\|EPI_ISL_451487\|2020- | .................... | .................... | ..................... | .................... | .................... | .................... |
| C | 7607 | hCoV-19/Australia/NSW222/2020\|EPI_ISL_451488\|2020- | .................... | .................... | ..................... | .................... | .................... | .................... |
| C | 7608 | hCoV-19/Australia/NSW223/2020\|EPI_ISL_451489\|2020- | .................... | .................... | ..................... | .................... | .................... | .................... |
| C | 7609 | hCoV-19/Australia/NSW224/2020\|EPI_ISL_451490\|2020- | .................... | .................... | ..................... | .................... | .................... | .................... |
| C | 7610 | hCoV-19/Australia/NSW225/2020\|EPI_ISL_451491\|2020- | .................... | .................... | ..................... | .................... | .................... | .................... |
| C | 7611 | hCoV-19/Australia/NSW226/2020\|EPI_ISL_451492\|2020- | .................... | .................... | ..................... | .................... | .................... | .................... |
| C | 7612 | hCoV-19/Australia/NSW227/2020\|EPI_ISL_451493\|2020- | .................... | .................... | ..................... | .................... | .................... | .................... |
| C | 7613 | hCoV-19/Australia/NSW228/2020\|EPI_ISL_451494\|2020- | .................... | .................... | ..................... | .................... | .................... | .................... |
| C | 7614 | hCoV-19/Australia/NSW229/2020\|EPI_ISL_451495\|2020- | .................... | .................... | ..................... | .................... | .................... | .................... |
| C | 7615 | hCoV-19/Australia/NSW230/2020\|EPI_ISL_451496\|2020- | .................... | .................... | ..................... | .................... | .................... | .................... |
| C | 7616 | hCoV-19/Australia/NSW231/2020\|EPI_ISL_451497\|2020- | .................... | .................... | ..................... | .................... | .................... | .................... |
| C | 7617 | hCoV-19/Australia/NSW232/2020\|EPI_ISL_451498\|2020- | .................... | .................... | ..................... | .................... | .................... | .................... |
| C | 7618 | hCoV-19/Australia/NSW233/2020\|EPI_ISL_451499\|2020- | .................... | .................... | ..................... | .................... | .................... | .................... |
| C | 7619 | hCoV-19/Spain/PV-IBV-000717/2020\|EPI_ISL_452751\|20 | .................... | .................... | ..................... | .................... | .................... | .................... |
| C | 7626 | hCoV-19/Spain/PV-IBV-000703/2020\|EPI_ISL_452763\|20 | .................... | .................... | ..................... | .................... | .................... | .................... |
| C | 7634 | hCoV-19/Spain/PV-IBV-000692/2020\|EPI_ISL_452772\|20 | .................... | .................... | ..................... | .................... | .................... | .................... |
| C | 7635 | hCoV-19/Spain/PV-IBV-000691/2020\|EPI_ISL_452773\|20 | .................... | .................... | ..................... | .................... | .................... | .................... |
| C | 7638 | hCoV-19/Spain/PV-IBV-000685/2020\|EPI_ISL_452777\|20 | .................... | .................... | ..................... | .................... | .................... | .................... |
| C | 7645 | hCoV-19/India/MP-NIHSAD-2604-22/2020\|EPI_ISL_45279 | .................... | .................... | ..................... | .................... | .................... | .................... |
| C | 7646 | hCoV-19/India/MP-NIHSAD-2904-107/2020\|EPI_ISL_4527 | .................... | .................... | ..................... | .................... | .................... | .................... |
| C | 7647 | hCoV-19/India/MP-NIHSAD-0105-77/2020\|EPI_ISL_45279 | .................... | .................... | ..................... | .................... | .................... | .................... |
| C | 7648 | hCoV-19/India/MP-NIHSAD-0405-24/2020\|EPI_ISL_45279 | .................... | .................... | ..................... | .................... | .................... | .................... |
| C | 7649 | hCoV-19/India/MP-NIHSAD-0905-216/2020\|EPI_ISL_4527 | .................... | .................... | ..................... | .................... | .................... | .................... |
| C | 7650 | hCoV-19/India/MP-NIHSAD-1005-52/2020\|EPI_ISL_45279 | .................... | .................... | ..................... | .................... | .................... | .................... |
| C | 7651 | hCoV-19/USA/VA-DCLS-0218/2020\|EPI_ISL_452796\|2020- | .................... | .................... | ..................... | .................... | .................... | .................... |
| C | 7652 | hCoV-19/USA/VA-DCLS-0219/2020\|EPI_ISL_452797\|2020- | .................... | .................... | ..................... | .................... | .................... | .................... |
| C | 7653 | hCoV-19/USA/VA-DCLS-0220/2020\|EPI_ISL_452798\|2020- | .................... | .................... | ..................... | .................... | .................... | .................... |
| C | 7654 | hCoV-19/New Zealand/20VR1899/2020\|EPI_ISL_456300\|2 | .................... | .................... | ..................... | .................... | .................... | .................... |
| C | 7655 | hCoV-19/England/SHEF-C3C50/2020\|EPI_ISL_457602\|202 | .................... | .................... | ..................... | .................... | .................... | .................... |
| C | 7656 | hCoV-19/England/SHEF-C42B7/2020\|EPI_ISL_457603\|202 | .................... | .................... | ..................... | .................... | .................... | .................... |
| C | 7657 | hCoV-19/England/SHEF-C42C6/2020\|EPI_ISL_457604\|202 | .................... | .................... | ..................... | .................... | .................... | .................... |
| C | 7658 | hCoV-19/England/SHEF-C469D/2020\|EPI_ISL_457605\|202 | .................... | .................... | ..................... | .................... | .................... | .................... |
| C | 7659 | hCoV-19/England/SHEF-C4F74/2020\|EPI_ISL_457606\|202 | .................... | .................... | ..................... | .................... | .................... | .................... |
| C | 7660 | hCoV-19/England/SHEF-C65AD/2020\|EPI_ISL_457611\|202 | .................... | .................... | ..................... | .................... | .................... | .................... |

| | | | | | | | |
|---|---|---|---|---|---|---|---|
| C | 7661 hCoV-19/England/SHEF-C7339/2020\|EPI_ISL_457617\|202 | ···················· | ···················· | ···················· | ···················· | ···················· | ···················· |
| C | 7662 hCoV-19/England/SHEF-C7454/2020\|EPI_ISL_457618\|202 | ···················· | ···················· | ···················· | ···················· | ···················· | ···················· |
| C | 7663 hCoV-19/England/SHEF-C7928/2020\|EPI_ISL_457620\|202 | ···················· | ···················· | ···················· | ···················· | ···················· | ···················· |
| C | 7664 hCoV-19/England/SHEF-C8857/2020\|EPI_ISL_457631\|202 | ···················· | ···················· | ···················· | ···················· | ···················· | ···················· |
| C | 7665 hCoV-19/England/BRIS-1262FE/2020\|EPI_ISL_458934\|20 | ···················· | ···················· | ···················· | ···················· | ···················· | ···················· |
| C | 7666 hCoV-19/England/BRIS-126E09/2020\|EPI_ISL_458937\|20 | ···················· | ···················· | ···················· | ···················· | ···················· | ···················· |
| C | 7667 hCoV-19/England/BRIS-126285/2020\|EPI_ISL_458938\|20 | ···················· | ···················· | ···················· | ···················· | ···················· | ···················· |
| C | 7668 hCoV-19/England/BRIS-126B4E/2020\|EPI_ISL_458939\|20 | ···················· | ·A·················· | ···················· | ···················· | ···················· | ···················· |
| C | 7669 hCoV-19/England/BRIS-126CF0/2020\|EPI_ISL_458941\|20 | ···················· | ···················· | ···················· | ···················· | ···················· | ···················· |
| C | 7670 hCoV-19/England/BRIS-12604F/2020\|EPI_ISL_458943\|20 | ···················· | ···················· | ···················· | ···················· | ···················· | ···················· |
| C | 7671 hCoV-19/England/BRIS-126443/2020\|EPI_ISL_458945\|20 | ···················· | ···················· | ···················· | ···················· | ···················· | ···················· |
| C | 7672 hCoV-19/England/BRIS-1265D7/2020\|EPI_ISL_458946\|20 | ···················· | ···················· | ···················· | ···················· | ···················· | ···················· |
| C | 7673 hCoV-19/England/BRIS-1264DA/2020\|EPI_ISL_458947\|20 | ···················· | ···················· | ···················· | ···················· | ···················· | ···················· |
| C | 7674 hCoV-19/England/BRIS-126768/2020\|EPI_ISL_458949\|20 | ···················· | ···················· | ···················· | ···················· | ···················· | ···················· |
| C | 7675 hCoV-19/England/BRIS-12673B/2020\|EPI_ISL_458950\|20 | ···················· | ···················· | ···················· | ···················· | ···················· | ···················· |
| C | 7676 hCoV-19/England/BRIS-1264E9/2020\|EPI_ISL_458951\|20 | ···················· | ···················· | ···················· | ···················· | ···················· | ···················· |
| C | 7677 hCoV-19/England/BRIS-12606D/2020\|EPI_ISL_458952\|20 | ···················· | ···················· | ···················· | ···················· | ···················· | ···················· |
| C | 7678 hCoV-19/England/BRIS-126962/2020\|EPI_ISL_458953\|20 | ···················· | ···················· | ···················· | ···················· | ···················· | ···················· |
| C | 7679 hCoV-19/England/BRIS-12681A/2020\|EPI_ISL_458962\|20 | ···················· | ···················· | ···················· | ···················· | ···················· | ···················· |
| C | 7680 hCoV-19/England/BRIS-1262C1/2020\|EPI_ISL_458963\|20 | ···················· | ···················· | ···················· | ···················· | ···················· | ···················· |
| C | 7681 hCoV-19/England/NORW-E8068/2020\|EPI_ISL_449100\|202 | ···················· | ···················· | ···················· | ···················· | ···················· | ···················· |
| C | 7682 hCoV-19/England/NORW-E8077/2020\|EPI_ISL_449101\|202 | ···················· | ···················· | ···················· | ···················· | ···················· | ···················· |
| C | 7683 hCoV-19/England/NORW-E8095/2020\|EPI_ISL_449102\|202 | ···················· | ···················· | ···················· | ···················· | ···················· | ···················· |
| C | 7684 hCoV-19/England/NORW-E80A4/2020\|EPI_ISL_449103\|202 | ···················· | ···················· | ···················· | ···················· | ···················· | ···················· |
| C | 7685 hCoV-19/England/NORW-E80B3/2020\|EPI_ISL_449104\|202 | ···················· | ···················· | ···················· | ···················· | ···················· | ···················· |
| C | 7686 hCoV-19/England/NORW-E80C2/2020\|EPI_ISL_449105\|202 | ···················· | ···················· | ···················· | ···················· | ···················· | ···················· |
| C | 7687 hCoV-19/England/NORW-E80D1/2020\|EPI_ISL_449106\|202 | ···················· | ···················· | ···················· | ···················· | ···················· | ···················· |
| C | 7688 hCoV-19/England/NORW-E80E0/2020\|EPI_ISL_449107\|202 | ···················· | ···················· | ···················· | ···················· | ···················· | ···················· |
| C | 7689 hCoV-19/England/NORW-E810B/2020\|EPI_ISL_449109\|202 | ···················· | ···················· | ···················· | ···················· | ···················· | ···················· |
| C | 7690 hCoV-19/England/NORW-E8578/2020\|EPI_ISL_449160\|202 | ···················· | ···················· | ···················· | ···················· | ···················· | ···················· |
| C | 7691 hCoV-19/England/NORW-E8587/2020\|EPI_ISL_449161\|202 | ···················· | ···················· | ···················· | ···················· | ···················· | ···················· |
| C | 7692 hCoV-19/England/NORW-E8596/2020\|EPI_ISL_449162\|202 | ···················· | ···················· | ···················· | ···················· | ···················· | ···················· |
| C | 7693 hCoV-19/England/NORW-E85B4/2020\|EPI_ISL_449163\|202 | ···················· | ···················· | ···················· | ···················· | ···················· | ···················· |
| C | 7694 hCoV-19/England/NORW-E85F0/2020\|EPI_ISL_449166\|202 | ···················· | ···················· | ···················· | ···················· | ···················· | ···················· |
| C | 7695 hCoV-19/USA/NY-PV09467/2020\|EPI_ISL_450140\|2020-03 | ···················· | ···················· | ···················· | ···················· | ···················· | ···················· |
| C | 7696 hCoV-19/USA/NY-PV09468/2020\|EPI_ISL_450141\|2020-03 | ···················· | ···················· | ···················· | ···················· | ···················· | ···················· |
| C | 7697 hCoV-19/USA/NY-PV09470/2020\|EPI_ISL_450142\|2020-03 | ···················· | ···················· | ···················· | ···················· | ···················· | ···················· |
| C | 7698 hCoV-19/USA/NY-PV09471/2020\|EPI_ISL_450143\|2020-03 | ···················· | ···················· | ···················· | ···················· | ···················· | ···················· |
| C | 7699 hCoV-19/USA/NY-NYUMC686/2020\|EPI_ISL_451401\|2020-0 | ···················· | ···················· | ···················· | ···················· | ···················· | ···················· |
| C | 7700 hCoV-19/USA/NY-NYUMC687/2020\|EPI_ISL_451402\|2020-0 | ···················· | ···················· | ···················· | ···················· | ···················· | ···················· |
| C | 7701 hCoV-19/USA/NY-NYUMC689/2020\|EPI_ISL_451404\|2020-0 | ···················· | ···················· | ···················· | ···················· | ···················· | ···················· |
| C | 7702 hCoV-19/USA/NY-NYUMC690/2020\|EPI_ISL_451405\|2020-0 | ···················· | ···················· | ···················· | ···················· | ···················· | ···················· |
| C | 7703 hCoV-19/USA/NY-NYUMC691/2020\|EPI_ISL_451406\|2020-0 | ···················· | ···················· | ···················· | ···················· | ···················· | ···················· |
| C | 7704 hCoV-19/USA/NY-NYUMC692/2020\|EPI_ISL_451407\|2020-0 | ···················· | ···················· | ···················· | ···················· | ···················· | ···················· |
| C | 7705 hCoV-19/USA/NY-NYUMC693/2020\|EPI_ISL_451408\|2020-0 | ···················· | ···················· | ···················· | ···················· | ···················· | ···················· |
| C | 7706 hCoV-19/USA/NY-NYUMC694/2020\|EPI_ISL_451409\|2020-0 | ···················· | ···················· | ···················· | ···················· | ···················· | ···················· |
| C | 7707 hCoV-19/USA/NY-NYUMC695/2020\|EPI_ISL_451410\|2020-0 | ···················· | ···················· | ···················· | ···················· | ···················· | ···················· |
| C | 7708 hCoV-19/USA/NY-NYUMC696/2020\|EPI_ISL_451411\|2020-0 | ···················· | ···················· | ···················· | ···················· | ···················· | ···················· |
| C | 7709 hCoV-19/USA/NY-NYUMC697/2020\|EPI_ISL_451412\|2020-0 | ···················· | ···················· | ···················· | ···················· | ···················· | ···················· |
| C | 7710 hCoV-19/USA/NY-NYUMC698/2020\|EPI_ISL_451413\|2020-0 | ···················· | ···················· | ···················· | ···················· | ···················· | ···················· |
| C | 7711 hCoV-19/USA/NY-NYUMC699/2020\|EPI_ISL_451414\|2020-0 | ···················· | ···················· | ···················· | ···················· | ···················· | ···················· |
| C | 7712 hCoV-19/USA/NY-NYUMC700/2020\|EPI_ISL_451415\|2020-0 | ···················· | ·············A······ | ···················· | ···················· | ···················· | ···················· |
| C | 7713 hCoV-19/USA/NY-NYUMC701/2020\|EPI_ISL_451416\|2020-0 | ···················· | ···················· | ···················· | ···················· | ···················· | ···················· |
| C | 7714 hCoV-19/USA/NY-NYUMC702/2020\|EPI_ISL_451417\|2020-0 | ···················· | ···················· | ···················· | ···················· | ···················· | ···················· |
| C | 7715 hCoV-19/USA/NY-NYUMC703/2020\|EPI_ISL_451418\|2020-0 | ···················· | ···················· | ···················· | ···················· | ···················· | ···················· |
| C | 7716 hCoV-19/USA/NY-NYUMC704/2020\|EPI_ISL_451419\|2020-0 | ···················· | ···················· | ···················· | ···················· | ···················· | ···················· |
| C | 7717 hCoV-19/USA/NY-NYUMC754/2020\|EPI_ISL_451468\|2020-0 | ···················· | ···················· | ···················· | ···················· | ···················· | ···················· |
| C | 7718 hCoV-19/USA/NY-NYUMC755/2020\|EPI_ISL_451469\|2020-0 | ···················· | ···················· | ···················· | ···················· | ···················· | ···················· |
| C | 7719 hCoV-19/USA/NY-NYUMC756/2020\|EPI_ISL_451470\|2020-0 | ···················· | ···················· | ···················· | ···················· | ···················· | ···················· |
| C | 7720 hCoV-19/USA/NY-NYUMC757/2020\|EPI_ISL_451471\|2020-0 | ···················· | ···················· | ···················· | ···················· | ···················· | ···················· |
| C | 7721 hCoV-19/USA/NY-NYUMC758/2020\|EPI_ISL_451472\|2020-0 | ···················· | ······A············· | ···················· | ···················· | ···················· | ···················· |
| C | 7722 hCoV-19/USA/NY-NYUMC760/2020\|EPI_ISL_451474\|2020-0 | ···················· | ···················· | ···················· | ···················· | ···················· | ···················· |
| C | 7724 hCoV-19/Spain/PV-IBV-000847/2020\|EPI_ISL_452701\|20 | ···················· | ···················· | ···················· | ···················· | ···················· | ···················· |
| C | 7725 hCoV-19/Spain/PV-IBV-000843/2020\|EPI_ISL_452703\|20 | ···················· | ···················· | ···················· | ···················· | ···················· | ···················· |
| C | 7735 hCoV-19/Spain/PV-IBV-000812/2020\|EPI_ISL_452715\|20 | ···················· | ···················· | ···················· | ···················· | ···················· | ···················· |
| C | 7737 hCoV-19/Spain/PV-IBV-000805/2020\|EPI_ISL_452717\|20 | ···················· | ···················· | ···················· | ···················· | ···················· | ···················· |
| C | 7738 hCoV-19/Spain/PV-IBV-000804/2020\|EPI_ISL_452718\|20 | ···················· | ···················· | ···················· | ···················· | ··NN················ | ···················· |
| C | 7741 hCoV-19/Spain/PV-IBV-000793/2020\|EPI_ISL_452721\|20 | ···················· | ···················· | ···················· | ···················· | ···················· | ···················· |
| C | 7742 hCoV-19/Spain/PV-IBV-000792/2020\|EPI_ISL_452722\|20 | ···················· | ···················· | ···················· | ···················· | ···················· | ···················· |
| C | 7746 hCoV-19/Spain/PV-IBV-000779/2020\|EPI_ISL_452726\|20 | ···················· | ···················· | ···················· | ···················· | ···················· | ···················· |
| C | 7754 hCoV-19/Spain/PV-IBV-000754/2020\|EPI_ISL_452735\|20 | ···················· | ···················· | ···················· | ···················· | ···················· | ···················· |
| C | 7757 hCoV-19/Spain/PV-IBV-000750/2020\|EPI_ISL_452738\|20 | ···················· | ···················· | ···················· | ···················· | ···················· | ···················· |
| C | 7761 hCoV-19/Spain/PV-IBV-000738/2020\|EPI_ISL_452742\|20 | ···················· | ···················· | ···················· | ···················· | ···················· | ···················· |
| C | 7763 hCoV-19/Spain/PV-IBV-000736/2020\|EPI_ISL_452744\|20 | ···················· | ···················· | ···················· | ···················· | ···················· | ···················· |
| C | 7766 hCoV-19/Spain/PV-IBV-000728/2020\|EPI_ISL_452747\|20 | ···················· | ···················· | ···················· | ···················· | ···················· | ···················· |
| C | 7767 hCoV-19/Spain/PV-IBV-000727/2020\|EPI_ISL_452748\|20 | ···················· | ···················· | ···················· | ···················· | ···················· | ···················· |
| C | 7768 hCoV-19/Spain/PV-IBV-000721/2020\|EPI_ISL_452749\|20 | ···················· | ···················· | ···················· | ···················· | ···················· | ···················· |
| C | 7770 hCoV-19/USA/VA-DCLS-0221/2020\|EPI_ISL_452799\|2020- | ···················· | ···················· | ···················· | ···················· | ···················· | ···················· |

| | | | | | | | | |
|---|---|---|---|---|---|---|---|---|
| C | 7771 | hCoV-19/New Zealand/20VR1275/2020\|EPI_ISL_456200\|2 | ···················· | ···················· | ···················· | ···················· | ···················· | ···················· |
| C | 7772 | hCoV-19/New Zealand/20VR1276/2020\|EPI_ISL_456201\|2 | ···················· | ···················· | ···················· | ···················· | ···················· | ···················· |
| C | 7773 | hCoV-19/New Zealand/20VR1277/2020\|EPI_ISL_456202\|2 | ···················· | ···················· | ···················· | ···················· | ···················· | ···················· |
| C | 7774 | hCoV-19/New Zealand/20VR1278/2020\|EPI_ISL_456203\|2 | ···················· | ···················· | ···················· | ···················· | ···················· | ···················· |
| C | 7775 | hCoV-19/New Zealand/20VR1280/2020\|EPI_ISL_456205\|2 | ···················· | ···················· | ···················· | ···················· | ···················· | ···················· |
| C | 7776 | hCoV-19/New Zealand/20VR1281/2020\|EPI_ISL_456206\|2 | ···················· | ···················· | ···················· | ···················· | ···················· | ···················· |
| C | 7777 | hCoV-19/New Zealand/20VR1286/2020\|EPI_ISL_456210\|2 | ···················· | ···················· | ···················· | ···················· | ···················· | ···················· |
| C | 7778 | hCoV-19/New Zealand/20VR1290/2020\|EPI_ISL_456212\|2 | ···················· | ···················· | ···················· | ···················· | ···················· | ···················· |
| C | 7779 | hCoV-19/New Zealand/20VR1297/2020\|EPI_ISL_456213\|2 | ···················· | ···················· | ···················· | ···················· | ···················· | ···················· |
| C | 7780 | hCoV-19/New Zealand/20VR1298/2020\|EPI_ISL_456214\|2 | ···················· | ···················· | ···················· | ···················· | ···················· | ···················· |
| C | 7781 | hCoV-19/New Zealand/20VR1380/2020\|EPI_ISL_456215\|2 | ···················· | ···················· | ···················· | ···················· | ···················· | ···················· |
| C | 7782 | hCoV-19/New Zealand/20VR1450/2020\|EPI_ISL_456217\|2 | ···················· | ···················· | ···················· | ···················· | ···················· | ···················· |
| C | 7783 | hCoV-19/New Zealand/20VR1451/2020\|EPI_ISL_456218\|2 | ···················· | ···················· | ···················· | ···················· | ···················· | ···················· |
| C | 7784 | hCoV-19/New Zealand/20VR1452/2020\|EPI_ISL_456219\|2 | ···················· | ·A·················· | ···················· | ···················· | ···················· | ···················· |
| C | 7785 | hCoV-19/New Zealand/20VR1454/2020\|EPI_ISL_456220\|2 | ···················· | ···················· | ···················· | ···················· | ···················· | ···················· |
| C | 7786 | hCoV-19/New Zealand/20VR1499/2020\|EPI_ISL_456221\|2 | ···················· | ···················· | ···················· | ···················· | ···················· | ···················· |
| C | 7787 | hCoV-19/New Zealand/20VR1561/2020\|EPI_ISL_456222\|2 | ···················· | ···················· | ···················· | ···················· | ···················· | ···················· |
| C | 7788 | hCoV-19/New Zealand/20VR1581/2020\|EPI_ISL_456223\|2 | ···················· | ···················· | ···················· | ···················· | ···················· | ···················· |
| C | 7789 | hCoV-19/New Zealand/20VR1597/2020\|EPI_ISL_456224\|2 | ···················· | ···················· | ···················· | ···················· | ···················· | ···················· |
| C | 7790 | hCoV-19/New Zealand/20VR1615/2020\|EPI_ISL_456225\|2 | ···················· | ···················· | ···················· | ···················· | ···················· | ···················· |
| C | 7791 | hCoV-19/New Zealand/20VR1618/2020\|EPI_ISL_456228\|2 | ···················· | ···················· | ···················· | ···················· | ···················· | ···················· |
| C | 7792 | hCoV-19/New Zealand/20VR1620/2020\|EPI_ISL_456230\|2 | ···················· | ···················· | ···················· | ···················· | ···················· | ···················· |
| C | 7793 | hCoV-19/New Zealand/20VR1623/2020\|EPI_ISL_456233\|2 | ···················· | ···················· | ···················· | ···················· | ···················· | ···················· |
| C | 7794 | hCoV-19/New Zealand/20VR1624/2020\|EPI_ISL_456234\|2 | ···················· | ···················· | ···················· | ···················· | ···················· | ···················· |
| C | 7795 | hCoV-19/New Zealand/20VR1626/2020\|EPI_ISL_456236\|2 | ···················· | ···················· | ···················· | ···················· | ···················· | ···················· |
| C | 7796 | hCoV-19/New Zealand/20VR1627/2020\|EPI_ISL_456237\|2 | ···················· | ···················· | ···················· | ···················· | ···················· | ···················· |
| C | 7797 | hCoV-19/New Zealand/20VR1756/2020\|EPI_ISL_456238\|2 | ···················· | ···················· | ···················· | ···················· | ···················· | ···················· |
| C | 7798 | hCoV-19/New Zealand/20VR1771/2020\|EPI_ISL_456242\|2 | ···················· | ···················· | ···················· | ···················· | ···················· | ···················· |
| C | 7799 | hCoV-19/New Zealand/20VR1772/2020\|EPI_ISL_456243\|2 | ···················· | ···················· | ···················· | ···················· | ···················· | ···················· |
| C | 7800 | hCoV-19/New Zealand/20VR1773/2020\|EPI_ISL_456244\|2 | ···················· | ···················· | ···················· | ···················· | ···················· | ···················· |
| C | 7801 | hCoV-19/New Zealand/20VR1776/2020\|EPI_ISL_456245\|2 | ···················· | ···················· | ···················· | ···················· | ···················· | ···················· |
| C | 7802 | hCoV-19/New Zealand/20VR1777/2020\|EPI_ISL_456246\|2 | ···················· | ···················· | ···················· | ···················· | ···················· | ···················· |
| C | 7803 | hCoV-19/New Zealand/20VR1778/2020\|EPI_ISL_456247\|2 | ···················· | ···················· | ···················· | ···················· | ···················· | ···················· |
| C | 7804 | hCoV-19/New Zealand/20VR1779/2020\|EPI_ISL_456248\|2 | ···················· | ···················· | ···················· | ···················· | ···················· | ···················· |
| C | 7805 | hCoV-19/New Zealand/20VR1780/2020\|EPI_ISL_456249\|2 | ···················· | ···················· | ···················· | ···················· | ···················· | ···················· |
| C | 7807 | hCoV-19/New Zealand/20VR1782/2020\|EPI_ISL_456251\|2 | ···················· | ···················· | ···················· | ···················· | ···················· | ···················· |
| C | 7808 | hCoV-19/New Zealand/20VR1783/2020\|EPI_ISL_456252\|2 | ···················· | ···················· | ···················· | ···················· | ···················· | ···················· |
| C | 7809 | hCoV-19/New Zealand/20VR1784/2020\|EPI_ISL_456253\|2 | ···················· | ···················· | ···················· | ···················· | ···················· | ···················· |
| C | 7810 | hCoV-19/New Zealand/20VR1785/2020\|EPI_ISL_456254\|2 | ···················· | ···················· | ···················· | ···················· | ···················· | ···················· |
| C | 7811 | hCoV-19/New Zealand/20VR1796/2020\|EPI_ISL_456255\|2 | ···················· | ···················· | ···················· | ···················· | ···················· | ···················· |
| C | 7812 | hCoV-19/New Zealand/20VR1797/2020\|EPI_ISL_456256\|2 | ···················· | ···················· | ···················· | ···················· | ···················· | ···················· |
| C | 7813 | hCoV-19/New Zealand/20VR1798/2020\|EPI_ISL_456257\|2 | ···················· | ···················· | ···················· | ···················· | ···················· | ···················· |
| C | 7814 | hCoV-19/New Zealand/20VR1799/2020\|EPI_ISL_456258\|2 | ···················· | ···················· | ···················· | ···················· | ···················· | ···················· |
| C | 7815 | hCoV-19/New Zealand/20VR1809/2020\|EPI_ISL_456261\|2 | ···················· | ···················· | ···················· | ···················· | ···················· | ···················· |
| C | 7816 | hCoV-19/New Zealand/20VR1814/2020\|EPI_ISL_456262\|2 | ···················· | ···················· | ···················· | ···················· | ···················· | ···················· |
| C | 7817 | hCoV-19/New Zealand/20VR1816/2020\|EPI_ISL_456263\|2 | ···················· | ···················· | ···················· | ···················· | ···················· | ···················· |
| C | 7818 | hCoV-19/New Zealand/20VR1817/2020\|EPI_ISL_456264\|2 | ···················· | ···················· | ···················· | ···················· | ···················· | ···················· |
| C | 7819 | hCoV-19/New Zealand/20VR1818/2020\|EPI_ISL_456265\|2 | ···················· | ···················· | ···················· | ···················· | ···················· | ···················· |
| C | 7820 | hCoV-19/New Zealand/20VR1821/2020\|EPI_ISL_456266\|2 | ···················· | ···················· | ···················· | ···················· | ···················· | ···················· |
| C | 7821 | hCoV-19/New Zealand/20VR1822/2020\|EPI_ISL_456267\|2 | ···················· | ···················· | ···················· | ···················· | ···················· | ···················· |
| C | 7822 | hCoV-19/New Zealand/20VR1823/2020\|EPI_ISL_456268\|2 | ···················· | ···················· | ···················· | ···················· | ···················· | ···················· |
| C | 7823 | hCoV-19/New Zealand/20VR1825/2020\|EPI_ISL_456270\|2 | ···················· | ···················· | ···················· | ···················· | ···················· | ···················· |
| C | 7824 | hCoV-19/New Zealand/20VR1826/2020\|EPI_ISL_456271\|2 | ···················· | ···················· | ···················· | ···················· | ···················· | ···················· |
| C | 7825 | hCoV-19/New Zealand/20VR1827/2020\|EPI_ISL_456272\|2 | ···················· | ···················· | ···················· | ···················· | ···················· | ···················· |
| C | 7826 | hCoV-19/New Zealand/20VR1828/2020\|EPI_ISL_456273\|2 | ···················· | ···················· | ···················· | ···················· | ···················· | ···················· |
| C | 7827 | hCoV-19/New Zealand/20VR1833/2020\|EPI_ISL_456276\|2 | ···················· | ···················· | ···················· | ···················· | ···················· | ···················· |
| C | 7828 | hCoV-19/New Zealand/20VR1837/2020\|EPI_ISL_456278\|2 | ···················· | ···················· | ···················· | ···················· | ···················· | ···················· |
| C | 7829 | hCoV-19/New Zealand/20VR1838/2020\|EPI_ISL_456279\|2 | ···················· | ···················· | ···················· | ···················· | ···················· | ···················· |
| C | 7830 | hCoV-19/New Zealand/20VR1839/2020\|EPI_ISL_456280\|2 | ···················· | ···················· | ···················· | ···················· | ···················· | ···················· |
| C | 7831 | hCoV-19/New Zealand/20VR1843/2020\|EPI_ISL_456281\|2 | ···················· | ···················· | ···················· | ···················· | ···················· | ···················· |
| C | 7832 | hCoV-19/New Zealand/20VR1845/2020\|EPI_ISL_456282\|2 | ···················· | ···················· | ···················· | ···················· | ···················· | ···················· |
| C | 7833 | hCoV-19/New Zealand/20VR1847/2020\|EPI_ISL_456283\|2 | ···················· | ···················· | ···················· | ···················· | ···················· | ···················· |
| C | 7834 | hCoV-19/New Zealand/20VR1856/2020\|EPI_ISL_456284\|2 | ···················· | ···················· | ···················· | ···················· | ···················· | ···················· |
| C | 7835 | hCoV-19/New Zealand/20VR1860/2020\|EPI_ISL_456287\|2 | ···················· | ···················· | ···················· | ···················· | ···················· | ···················· |
| C | 7836 | hCoV-19/New Zealand/20VR1862/2020\|EPI_ISL_456288\|2 | ···················· | ···················· | ···················· | ···················· | ···················· | ···················· |
| C | 7837 | hCoV-19/New Zealand/20VR1863/2020\|EPI_ISL_456289\|2 | ···················· | ···················· | ···················· | ···················· | ···················· | ···················· |
| C | 7838 | hCoV-19/New Zealand/20VR1864/2020\|EPI_ISL_456290\|2 | ···················· | ···················· | ···················· | ···················· | ···················· | ···················· |
| C | 7839 | hCoV-19/New Zealand/20VR1867/2020\|EPI_ISL_456292\|2 | ···················· | ···················· | ···················· | ···················· | ···················· | ···················· |
| C | 7840 | hCoV-19/New Zealand/20VR1887/2020\|EPI_ISL_456294\|2 | ···················· | ···················· | ···················· | ···················· | ···················· | ···················· |
| C | 7841 | hCoV-19/New Zealand/20VR1890/2020\|EPI_ISL_456295\|2 | ···················· | ···················· | ···················· | ···················· | ···················· | ···················· |
| C | 7842 | hCoV-19/New Zealand/20VR1891/2020\|EPI_ISL_456296\|2 | ···················· | ···················· | ···················· | ···················· | ···················· | ···················· |
| C | 7843 | hCoV-19/New Zealand/20VR1893/2020\|EPI_ISL_456297\|2 | ···················· | ···················· | ···················· | ···················· | ···················· | ···················· |
| C | 7844 | hCoV-19/New Zealand/20VR1897/2020\|EPI_ISL_456299\|2 | ···················· | ···················· | ···················· | ···················· | ···················· | ···················· |
| C | 7846 | hCoV-19/England/NORW-EC42C/2020\|EPI_ISL_457525\|202 | ···················· | ···················· | ···················· | ···················· | ···················· | ···················· |
| C | 7847 | hCoV-19/England/NORW-EC4A4/2020\|EPI_ISL_457527\|202 | ···················· | ···················· | ···················· | ···················· | ···················· | ···················· |
| C | 7848 | hCoV-19/England/NORW-EC4C2/2020\|EPI_ISL_457528\|202 | ···················· | ···················· | ···················· | ···················· | ···················· | ···················· |
| C | 7849 | hCoV-19/England/NORW-EC4D1/2020\|EPI_ISL_457529\|202 | ···················· | ···················· | ···················· | ···················· | ···················· | ···················· |
| C | 7850 | hCoV-19/England/NORW-EC4E0/2020\|EPI_ISL_457530\|202 | ···················· | ···················· | ···················· | ···················· | ···················· | ···················· |

| | | | | | | | | |
|---|---|---|---|---|---|---|---|---|
| C | 7851 hCoV-19/England/NORW-EC4FF/2020\|EPI_ISL_457531\|202 | .................... | .................... | .................... | .................... | .................... | .................... | |
| C | 7852 hCoV-19/England/NORW-EC50B/2020\|EPI_ISL_457532\|202 | .................... | ...................A | .................... | .................... | .................... | .................... | 1 |
| C | 7853 hCoV-19/England/NORW-EC529/2020\|EPI_ISL_457533\|202 | .................... | .................... | .................... | .................... | .................... | .................... | |
| C | 7854 hCoV-19/England/NORW-EC592/2020\|EPI_ISL_457534\|202 | .................... | .................... | .................... | .................... | .................... | .................... | |
| C | 7855 hCoV-19/England/NORW-EC5A1/2020\|EPI_ISL_457535\|202 | .................... | .................... | .................... | .................... | .................... | .................... | |
| C | 7856 hCoV-19/England/NORW-EC5DE/2020\|EPI_ISL_457538\|202 | .................... | .................... | .................... | .................... | .................... | .................... | |
| C | 7857 hCoV-19/England/NORW-EC5ED/2020\|EPI_ISL_457539\|202 | .................... | .................... | .................... | .................... | .................... | .................... | |
| C | 7858 hCoV-19/England/NORW-EC5FC/2020\|EPI_ISL_457540\|202 | .................... | .................... | .................... | .................... | .................... | .................... | |
| C | 7859 hCoV-19/England/NORW-EC617/2020\|EPI_ISL_457541\|202 | .................... | .................... | .................... | .................... | .................... | .................... | |
| C | 7860 hCoV-19/England/NORW-EC626/2020\|EPI_ISL_457542\|202 | .................... | .................... | .................... | .................... | .................... | .................... | |
| C | 7861 hCoV-19/England/NORW-EC644/2020\|EPI_ISL_457543\|202 | .................... | .................... | .................... | .................... | .................... | .................... | |
| C | 7862 hCoV-19/England/NORW-EC653/2020\|EPI_ISL_457544\|202 | .................... | .................... | .................... | .................... | .................... | .................... | |
| C | 7863 hCoV-19/England/NORW-EC662/2020\|EPI_ISL_457545\|202 | .................... | ...................A | .................... | .................... | .................... | .................... | 1 |
| C | 7864 hCoV-19/England/NORW-EC671/2020\|EPI_ISL_457546\|202 | .................... | .................... | .................... | .................... | .................... | .................... | |
| C | 7865 hCoV-19/England/NORW-EC680/2020\|EPI_ISL_457547\|202 | .................... | .................... | .................... | .................... | .................... | .................... | |
| C | 7866 hCoV-19/England/NORW-EC69F/2020\|EPI_ISL_457548\|202 | .................... | ...................A | .................... | .................... | .................... | .................... | 1 |
| C | 7867 hCoV-19/England/NORW-EC6BD/2020\|EPI_ISL_457550\|202 | .................... | .................... | .................... | .................... | .................... | .................... | |
| C | 7868 hCoV-19/England/NORW-EC6DB/2020\|EPI_ISL_457551\|202 | .................... | .................... | .................... | .................... | .................... | .................... | |
| C | 7869 hCoV-19/England/NORW-EC6EA/2020\|EPI_ISL_457552\|202 | .................... | .................... | .................... | .................... | .................... | .................... | |
| C | 7870 hCoV-19/England/NORW-EC6F9/2020\|EPI_ISL_457553\|202 | .................... | .................... | .................... | .................... | .................... | .................... | |
| C | 7871 hCoV-19/England/NORW-EC714/2020\|EPI_ISL_457555\|202 | .................... | ...................A | .................... | .................... | .................... | .................... | 1 |
| C | 7872 hCoV-19/England/NORW-EC732/2020\|EPI_ISL_457556\|202 | .................... | .................... | .................... | .................... | .................... | .................... | |
| C | 7873 hCoV-19/England/NORW-EC750/2020\|EPI_ISL_457558\|202 | .................... | .................... | .................... | .................... | .................... | .................... | |
| C | 7874 hCoV-19/England/NORW-EC76F/2020\|EPI_ISL_457559\|202 | .................... | ...................A | .................... | .................... | .................... | .................... | 1 |
| C | 7875 hCoV-19/England/NORW-EC77E/2020\|EPI_ISL_457560\|202 | .................... | .................... | .................... | .................... | .................... | .................... | |
| C | 7876 hCoV-19/England/NORW-EC78D/2020\|EPI_ISL_457561\|202 | .................... | .................... | .................... | .................... | .................... | .................... | |
| C | 7877 hCoV-19/England/NORW-EC7AB/2020\|EPI_ISL_457562\|202 | .................... | .................... | .................... | .................... | .................... | .................... | |
| C | 7878 hCoV-19/England/NORW-EC7E7/2020\|EPI_ISL_457563\|202 | .................... | .................... | .................... | .................... | .................... | .................... | |
| C | 7879 hCoV-19/England/NORW-EC802/2020\|EPI_ISL_457565\|202 | .................... | .................... | .................... | .................... | .................... | .................... | |
| C | 7880 hCoV-19/England/NORW-EC85D/2020\|EPI_ISL_457566\|202 | .................... | .................... | .................... | .................... | .................... | .................... | |
| C | 7881 hCoV-19/England/NORW-EC87B/2020\|EPI_ISL_457567\|202 | .................... | .................... | .................... | .................... | .................... | .................... | |
| C | 7882 hCoV-19/England/NORW-EC899/2020\|EPI_ISL_457568\|202 | .................... | .................... | .................... | .................... | .................... | .................... | |
| C | 7883 hCoV-19/England/NORW-EC8B7/2020\|EPI_ISL_457569\|202 | .................... | .................... | .................... | .................... | .................... | .................... | |
| C | 7884 hCoV-19/England/NORW-EC8C6/2020\|EPI_ISL_457570\|202 | .................... | .................... | .................... | .................... | .................... | .................... | |
| C | 7885 hCoV-19/England/NORW-EC8E4/2020\|EPI_ISL_457571\|202 | .................... | .................... | .................... | .................... | .................... | .................... | |
| C | 7886 hCoV-19/England/NORW-EC969/2020\|EPI_ISL_457572\|202 | .................... | .................... | .................... | .................... | .................... | .................... | |
| C | 7887 hCoV-19/England/NOTT-1114D2/2020\|EPI_ISL_457574\|20 | .................... | .................... | .................... | .................... | .................... | .................... | |
| C | 7888 hCoV-19/England/NOTT-1114F0/2020\|EPI_ISL_457575\|20 | .................... | .................... | .................... | .................... | .................... | .................... | |
| C | 7889 hCoV-19/England/NOTT-111539/2020\|EPI_ISL_457577\|20 | .................... | .................... | .................... | .................... | .................... | .................... | |
| C | 7890 hCoV-19/England/NOTT-111548/2020\|EPI_ISL_457578\|20 | .................... | .................... | .................... | .................... | .................... | .................... | |
| C | 7891 hCoV-19/England/NOTT-111557/2020\|EPI_ISL_457579\|20 | .................... | .................... | .................... | .................... | .................... | .................... | |
| C | 7892 hCoV-19/England/SHEF-C0989/2020\|EPI_ISL_457581\|202 | .................... | .................... | .................... | .................... | .................... | .................... | |
| C | 7893 hCoV-19/England/SHEF-C0A68/2020\|EPI_ISL_457582\|202 | .................... | .................... | .................... | .................... | .................... | .................... | |
| C | 7894 hCoV-19/England/SHEF-C0C08/2020\|EPI_ISL_457583\|202 | .................... | .................... | .................... | .................... | .................... | .................... | |
| C | 7895 hCoV-19/England/SHEF-C1029/2020\|EPI_ISL_457584\|202 | .................... | .................... | .................... | .................... | .................... | .................... | |
| C | 7896 hCoV-19/England/SHEF-C179D/2020\|EPI_ISL_457586\|202 | .................... | .................... | .................... | .................... | .................... | .................... | |
| C | 7897 hCoV-19/England/SHEF-C2477/2020\|EPI_ISL_457592\|202 | .................... | .................... | .................... | .................... | .................... | .................... | |
| C | 7898 hCoV-19/England/SHEF-C2671/2020\|EPI_ISL_457593\|202 | .................... | .................... | .................... | .................... | .................... | .................... | |
| C | 7899 hCoV-19/England/SHEF-C27F6/2020\|EPI_ISL_457594\|202 | .................... | .................... | .................... | .................... | .................... | .................... | |
| C | 7900 hCoV-19/England/SHEF-C2C7F/2020\|EPI_ISL_457596\|202 | .................... | .................... | .................... | .................... | .................... | .................... | |
| C | 7901 hCoV-19/England/SHEF-C3063/2020\|EPI_ISL_457597\|202 | .................... | .................... | .................... | .................... | .................... | .................... | |
| C | 7902 hCoV-19/England/SHEF-C33D3/2020\|EPI_ISL_457598\|202 | .................... | .................... | .................... | .................... | .................... | .................... | |
| C | 7903 hCoV-19/England/SHEF-C35DD/2020\|EPI_ISL_457599\|202 | .................... | .................... | .................... | .................... | .................... | .................... | |
| C | 7904 hCoV-19/Italy/TAA-AMVRC-2629/2020\|EPI_ISL_457700\|2 | .................... | .................... | .................... | .................... | .................... | .................... | |
| C | 7905 hCoV-19/Oman/RESP-20-797/2020\|EPI_ISL_457701\|2020- | .................... | .................... | .................... | .................... | .................... | .................... | |
| C | 7906 hCoV-19/Oman/205028718/2020\|EPI_ISL_457702\|2020-04 | .................... | .................... | .................... | .................... | .................... | .................... | |
| C | 7907 hCoV-19/Oman/205024123/2020\|EPI_ISL_457703\|2020-03 | .................... | .................... | .................... | .................... | .................... | .................... | |
| C | 7908 hCoV-19/Oman/RESP-20-837/2020\|EPI_ISL_457704\|2020- | .................... | .................... | .................... | .................... | .................... | .................... | |
| C | 7909 hCoV-19/Oman/205034466/2020\|EPI_ISL_457705\|2020-05 | .................... | .................... | .................... | .................... | .................... | .................... | |
| C | 7910 hCoV-19/Oman/RESP-20-1189/2020\|EPI_ISL_457706\|2020 | .................... | .................... | .................... | .................... | .................... | .................... | |
| C | 7911 hCoV-19/Oman/205024076/2020\|EPI_ISL_457707\|2020-03 | .................... | .................... | .................... | .................... | .................... | .................... | |
| C | 7912 hCoV-19/Estonia/ChVir4261/2020\|EPI_ISL_457719\|2020 | .................... | .................... | .................... | .................... | .................... | .................... | |
| C | 7913 hCoV-19/Italy/SIC-AMVRC-4411/2020\|EPI_ISL_457721\|2 | .................... | .................... | .................... | .................... | .................... | .................... | |
| C | 7914 hCoV-19/Estonia/ChVir4266/2020\|EPI_ISL_457722\|2020 | .................... | .................... | .................... | .................... | .................... | .................... | |
| C | 7916 hCoV-19/England/BRIS-1291A3/2020\|EPI_ISL_458856\|20 | .................... | .................... | .................... | .................... | .................... | .................... | |
| C | 7917 hCoV-19/England/BRIS-1290C4/2020\|EPI_ISL_458858\|20 | .................... | .................... | .................... | .................... | .................... | .................... | |
| C | 7918 hCoV-19/England/BRIS-128E43/2020\|EPI_ISL_458859\|20 | .................... | .................... | .................... | .................... | .................... | .................... | |
| C | 7919 hCoV-19/England/BRIS-1293F8/2020\|EPI_ISL_458861\|20 | .................... | .................... | .................... | .................... | .................... | .................... | |
| C | 7920 hCoV-19/England/BRIS-129747/2020\|EPI_ISL_458862\|20 | .................... | .................... | .................... | .................... | .................... | .................... | |
| C | 7921 hCoV-19/England/BRIS-129316/2020\|EPI_ISL_458863\|20 | .................... | .............A...... | .................... | .................... | .................... | .................... | |
| C | 7922 hCoV-19/England/BRIS-129589/2020\|EPI_ISL_458864\|20 | .................... | .................... | .................... | .................... | .................... | .................... | |
| C | 7923 hCoV-19/England/BRIS-128D0A/2020\|EPI_ISL_458866\|20 | .................... | .................... | .................... | .................... | .................... | .................... | |
| C | 7924 hCoV-19/England/BRIS-128C67/2020\|EPI_ISL_458868\|20 | .................... | .................... | .................... | .................... | .................... | .................... | |
| C | 7925 hCoV-19/England/BRIS-128CC1/2020\|EPI_ISL_458871\|20 | .................... | .................... | .................... | .................... | .................... | .................... | |
| C | 7926 hCoV-19/England/BRIS-128CA3/2020\|EPI_ISL_458877\|20 | .................... | .................... | .................... | .................... | .................... | .................... | |
| C | 7927 hCoV-19/England/BRIS-128827/2020\|EPI_ISL_458878\|20 | .................... | .A.................. | .................... | .................... | .................... | .................... | |
| C | 7928 hCoV-19/England/BRIS-1289E8/2020\|EPI_ISL_458879\|20 | .................... | .................... | .................... | .................... | .................... | .................... | |
| C | 7929 hCoV-19/England/BRIS-1286F0/2020\|EPI_ISL_458880\|20 | .................... | .................... | .................... | .................... | .................... | .................... | |

<table>
<tr><td>C</td><td>7930</td><td>hCoV-19/England/BRIS-128BB5/2020|EPI_ISL_458883|20</td><td>····················</td><td>····················</td><td>····················</td><td>····················</td><td>····················</td><td>····················</td><td></td><td></td><td></td></tr>
<tr><td>C</td><td>7931</td><td>hCoV-19/England/BRIS-129194/2020|EPI_ISL_458885|20</td><td>····················</td><td>····················</td><td>····················</td><td>····················</td><td>····················</td><td>····················</td><td></td><td></td><td></td></tr>
<tr><td>C</td><td>7932</td><td>hCoV-19/England/BRIS-128B00/2020|EPI_ISL_458886|20</td><td>····················</td><td>····················</td><td>····················</td><td>····················</td><td>····················</td><td>····················</td><td></td><td></td><td></td></tr>
<tr><td>C</td><td>7933</td><td>hCoV-19/England/BRIS-1286B4/2020|EPI_ISL_458888|20</td><td>····················</td><td>··---···············</td><td>····················</td><td>····················</td><td>····················</td><td>····················</td><td></td><td>1</td><td></td></tr>
<tr><td>C</td><td>7934</td><td>hCoV-19/England/BRIS-128E34/2020|EPI_ISL_458892|20</td><td>····················</td><td>····················</td><td>····················</td><td>····················</td><td>····················</td><td>····················</td><td></td><td></td><td></td></tr>
<tr><td>C</td><td>7935</td><td>hCoV-19/England/BRIS-12964A/2020|EPI_ISL_458894|20</td><td>····················</td><td>·············A······</td><td>····················</td><td>····················</td><td>····················</td><td>····················</td><td></td><td></td><td></td></tr>
<tr><td>C</td><td>7936</td><td>hCoV-19/England/BRIS-1297A1/2020|EPI_ISL_458899|20</td><td>····················</td><td>····················</td><td>····················</td><td>····················</td><td>····················</td><td>····················</td><td></td><td></td><td></td></tr>
<tr><td>C</td><td>7937</td><td>hCoV-19/Scotland/CVR3211/2020|EPI_ISL_449230|2020-</td><td>····················</td><td>····················</td><td>····················</td><td>····················</td><td>····················</td><td>····················</td><td></td><td></td><td></td></tr>
<tr><td>C</td><td>7938</td><td>hCoV-19/Scotland/CVR3212/2020|EPI_ISL_449231|2020-</td><td>····················</td><td>····················</td><td>····················</td><td>····················</td><td>····················</td><td>····················</td><td></td><td></td><td></td></tr>
<tr><td>C</td><td>7939</td><td>hCoV-19/Scotland/CVR3215/2020|EPI_ISL_449232|2020-</td><td>····················</td><td>···················A</td><td>····················</td><td>····················</td><td>····················</td><td>····················</td><td>1</td><td></td><td></td></tr>
<tr><td>C</td><td>7940</td><td>hCoV-19/Scotland/CVR3216/2020|EPI_ISL_449233|2020-</td><td>····················</td><td>····················</td><td>····················</td><td>····················</td><td>····················</td><td>····················</td><td></td><td></td><td></td></tr>
<tr><td>C</td><td>7941</td><td>hCoV-19/Scotland/CVR3217/2020|EPI_ISL_449234|2020-</td><td>····················</td><td>···················A</td><td>····················</td><td>····················</td><td>····················</td><td>····················</td><td>1</td><td></td><td></td></tr>
<tr><td>C</td><td>7942</td><td>hCoV-19/Scotland/CVR3221/2020|EPI_ISL_449235|2020-</td><td>····················</td><td>····················</td><td>····················</td><td>····················</td><td>····················</td><td>····················</td><td></td><td></td><td></td></tr>
<tr><td>C</td><td>7943</td><td>hCoV-19/Scotland/CVR3222/2020|EPI_ISL_449236|2020-</td><td>····················</td><td>····················</td><td>····················</td><td>····················</td><td>····················</td><td>····················</td><td></td><td></td><td></td></tr>
<tr><td>C</td><td>7944</td><td>hCoV-19/Scotland/CVR3225/2020|EPI_ISL_449237|2020-</td><td>····················</td><td>····················</td><td>····················</td><td>····················</td><td>····················</td><td>····················</td><td></td><td></td><td></td></tr>
<tr><td>C</td><td>7945</td><td>hCoV-19/Scotland/CVR3227/2020|EPI_ISL_449238|2020-</td><td>····················</td><td>····················</td><td>····················</td><td>····················</td><td>····················</td><td>····················</td><td></td><td></td><td></td></tr>
<tr><td>C</td><td>7946</td><td>hCoV-19/Scotland/CVR3230/2020|EPI_ISL_449240|2020-</td><td>····················</td><td>····················</td><td>····················</td><td>····················</td><td>····················</td><td>····················</td><td></td><td></td><td></td></tr>
<tr><td>C</td><td>7947</td><td>hCoV-19/Scotland/CVR3231/2020|EPI_ISL_449241|2020-</td><td>····················</td><td>····················</td><td>····················</td><td>····················</td><td>····················</td><td>····················</td><td></td><td></td><td></td></tr>
<tr><td>C</td><td>7948</td><td>hCoV-19/Scotland/CVR3232/2020|EPI_ISL_449242|2020-</td><td>····················</td><td>···················A</td><td>····················</td><td>····················</td><td>····················</td><td>····················</td><td>1</td><td></td><td></td></tr>
<tr><td>C</td><td>7949</td><td>hCoV-19/Scotland/CVR3233/2020|EPI_ISL_449243|2020-</td><td>····················</td><td>···················A</td><td>····················</td><td>····················</td><td>····················</td><td>····················</td><td>1</td><td></td><td></td></tr>
<tr><td>C</td><td>7950</td><td>hCoV-19/Scotland/CVR3234/2020|EPI_ISL_449244|2020-</td><td>····················</td><td>···················A</td><td>····················</td><td>····················</td><td>····················</td><td>····················</td><td>1</td><td></td><td></td></tr>
<tr><td>C</td><td>7951</td><td>hCoV-19/Scotland/CVR3235/2020|EPI_ISL_449245|2020-</td><td>····················</td><td>···················A</td><td>····················</td><td>····················</td><td>····················</td><td>····················</td><td>1</td><td></td><td></td></tr>
<tr><td>C</td><td>7952</td><td>hCoV-19/Scotland/CVR3236/2020|EPI_ISL_449246|2020-</td><td>····················</td><td>····················</td><td>····················</td><td>····················</td><td>····················</td><td>····················</td><td></td><td></td><td></td></tr>
<tr><td>C</td><td>7953</td><td>hCoV-19/Scotland/CVR3237/2020|EPI_ISL_449247|2020-</td><td>····················</td><td>···················A</td><td>····················</td><td>····················</td><td>····················</td><td>····················</td><td>1</td><td></td><td></td></tr>
<tr><td>C</td><td>7954</td><td>hCoV-19/Scotland/CVR3238/2020|EPI_ISL_449248|2020-</td><td>····················</td><td>·A··················</td><td>····················</td><td>····················</td><td>····················</td><td>····················</td><td></td><td></td><td></td></tr>
<tr><td>C</td><td>7955</td><td>hCoV-19/Scotland/CVR3239/2020|EPI_ISL_449249|2020-</td><td>····················</td><td>···················A</td><td>····················</td><td>····················</td><td>····················</td><td>····················</td><td>1</td><td></td><td></td></tr>
<tr><td>C</td><td>7956</td><td>hCoV-19/Scotland/CVR3242/2020|EPI_ISL_449250|2020-</td><td>····················</td><td>···················A</td><td>····················</td><td>····················</td><td>····················</td><td>····················</td><td>1</td><td></td><td></td></tr>
<tr><td>C</td><td>7957</td><td>hCoV-19/Scotland/CVR3243/2020|EPI_ISL_449251|2020-</td><td>····················</td><td>····················</td><td>····················</td><td>····················</td><td>····················</td><td>····················</td><td></td><td></td><td></td></tr>
<tr><td>C</td><td>7958</td><td>hCoV-19/Scotland/CVR3244/2020|EPI_ISL_449252|2020-</td><td>····················</td><td>····················</td><td>····················</td><td>····················</td><td>····················</td><td>····················</td><td></td><td></td><td></td></tr>
<tr><td>C</td><td>7959</td><td>hCoV-19/Scotland/CVR3245/2020|EPI_ISL_449253|2020-</td><td>····················</td><td>··········A·········</td><td>····················</td><td>····················</td><td>····················</td><td>····················</td><td></td><td></td><td></td></tr>
<tr><td>C</td><td>7960</td><td>hCoV-19/Scotland/CVR3246/2020|EPI_ISL_449254|2020-</td><td>····················</td><td>·A··················</td><td>····················</td><td>····················</td><td>····················</td><td>····················</td><td></td><td></td><td></td></tr>
<tr><td>C</td><td>7961</td><td>hCoV-19/Scotland/CVR3248/2020|EPI_ISL_449255|2020-</td><td>····················</td><td>····················</td><td>····················</td><td>····················</td><td>····················</td><td>····················</td><td></td><td></td><td></td></tr>
<tr><td>C</td><td>7962</td><td>hCoV-19/Scotland/CVR3249/2020|EPI_ISL_449256|2020-</td><td>····················</td><td>····················</td><td>····················</td><td>····················</td><td>····················</td><td>····················</td><td></td><td></td><td></td></tr>
<tr><td>C</td><td>7963</td><td>hCoV-19/Scotland/CVR3250/2020|EPI_ISL_449257|2020-</td><td>····················</td><td>····················</td><td>····················</td><td>····················</td><td>····················</td><td>····················</td><td></td><td></td><td></td></tr>
<tr><td>C</td><td>7964</td><td>hCoV-19/Scotland/CVR3251/2020|EPI_ISL_449258|2020-</td><td>····················</td><td>····················</td><td>····················</td><td>····················</td><td>····················</td><td>····················</td><td></td><td></td><td></td></tr>
<tr><td>C</td><td>7972</td><td>hCoV-19/Scotland/EDB4154/2020|EPI_ISL_449275|2020-</td><td>····················</td><td>···················A</td><td>····················</td><td>····················</td><td>····················</td><td>····················</td><td>1</td><td></td><td></td></tr>
<tr><td>C</td><td>7973</td><td>hCoV-19/Scotland/EDB4155/2020|EPI_ISL_449276|2020-</td><td>····················</td><td>···················A</td><td>····················</td><td>····················</td><td>····················</td><td>····················</td><td>1</td><td></td><td></td></tr>
<tr><td>C</td><td>7974</td><td>hCoV-19/Scotland/EDB4170/2020|EPI_ISL_449277|2020-</td><td>····················</td><td>···················A</td><td>····················</td><td>····················</td><td>····················</td><td>····················</td><td>1</td><td></td><td></td></tr>
<tr><td>C</td><td>7975</td><td>hCoV-19/Scotland/EDB4172/2020|EPI_ISL_449278|2020-</td><td>····················</td><td>···················A</td><td>····················</td><td>····················</td><td>····················</td><td>····················</td><td>1</td><td></td><td></td></tr>
<tr><td>C</td><td>7977</td><td>hCoV-19/Scotland/EDB4189/2020|EPI_ISL_449280|2020-</td><td>····················</td><td>···················A</td><td>····················</td><td>····················</td><td>····················</td><td>····················</td><td>1</td><td></td><td></td></tr>
<tr><td>C</td><td>7979</td><td>hCoV-19/Scotland/EDB4204/2020|EPI_ISL_449282|2020-</td><td>····················</td><td>···················A</td><td>····················</td><td>····················</td><td>····················</td><td>····················</td><td>1</td><td></td><td></td></tr>
<tr><td>C</td><td>7981</td><td>hCoV-19/Scotland/EDB4223/2020|EPI_ISL_449284|2020-</td><td>····················</td><td>···················A</td><td>····················</td><td>····················</td><td>····················</td><td>····················</td><td>1</td><td></td><td></td></tr>
<tr><td>C</td><td>7982</td><td>hCoV-19/Scotland/EDB4254/2020|EPI_ISL_449286|2020-</td><td>····················</td><td>···················A</td><td>····················</td><td>····················</td><td>····················</td><td>····················</td><td>1</td><td></td><td></td></tr>
<tr><td>C</td><td>7983</td><td>hCoV-19/Scotland/EDB4271/2020|EPI_ISL_449288|2020-</td><td>····················</td><td>···················A</td><td>····················</td><td>····················</td><td>····················</td><td>····················</td><td>1</td><td></td><td></td></tr>
<tr><td>C</td><td>7985</td><td>hCoV-19/Scotland/EDB4295/2020|EPI_ISL_449290|2020-</td><td>····················</td><td>···················A</td><td>····················</td><td>····················</td><td>····················</td><td>····················</td><td>1</td><td></td><td></td></tr>
<tr><td>C</td><td>7986</td><td>hCoV-19/Scotland/EDB4300/2020|EPI_ISL_449291|2020-</td><td>····················</td><td>···················A</td><td>····················</td><td>····················</td><td>····················</td><td>····················</td><td>1</td><td></td><td></td></tr>
<tr><td>C</td><td>7989</td><td>hCoV-19/Scotland/EDB4324/2020|EPI_ISL_449294|2020-</td><td>····················</td><td>···················A</td><td>····················</td><td>····················</td><td>····················</td><td>NN·N················</td><td>1</td><td>1</td><td>1</td></tr>
<tr><td>C</td><td>7992</td><td>hCoV-19/Scotland/EDB4362/2020|EPI_ISL_449297|2020-</td><td>····················</td><td>···················A</td><td>····················</td><td>····················</td><td>····················</td><td>····················</td><td>1</td><td></td><td></td></tr>
<tr><td>C</td><td>7993</td><td>hCoV-19/Scotland/EDB4365/2020|EPI_ISL_449298|2020-</td><td>····················</td><td>····················</td><td>····················</td><td>····················</td><td>····················</td><td>····················</td><td></td><td></td><td></td></tr>
<tr><td>C</td><td>7994</td><td>hCoV-19/Scotland/EDB4366/2020|EPI_ISL_449299|2020-</td><td>····················</td><td>····················</td><td>····················</td><td>····················</td><td>····················</td><td>····················</td><td></td><td></td><td></td></tr>
<tr><td>C</td><td>7995</td><td>hCoV-19/USA/LA-SR0308/2020|EPI_ISL_450231|2020-03-</td><td>····················</td><td>····················</td><td>····················</td><td>····················</td><td>····················</td><td>····················</td><td></td><td></td><td></td></tr>
<tr><td>C</td><td>7996</td><td>hCoV-19/USA/CA-UCSF-UC40/2020|EPI_ISL_450232|2020-</td><td>····················</td><td>····················</td><td>····················</td><td>····················</td><td>····················</td><td>····················</td><td></td><td></td><td></td></tr>
<tr><td>C</td><td>7997</td><td>hCoV-19/USA/CA-UCSF-UC41/2020|EPI_ISL_450233|2020-</td><td>····················</td><td>····················</td><td>····················</td><td>····················</td><td>····················</td><td>····················</td><td></td><td></td><td></td></tr>
<tr><td>C</td><td>7998</td><td>hCoV-19/USA/CA-UCSF-UC43/2020|EPI_ISL_450235|2020-</td><td>····················</td><td>····················</td><td>····················</td><td>····················</td><td>····················</td><td>····················</td><td></td><td></td><td></td></tr>
<tr><td>C</td><td>7999</td><td>hCoV-19/USA/CA-UCSF-UC44/2020|EPI_ISL_450236|2020-</td><td>····················</td><td>····················</td><td>····················</td><td>····················</td><td>····················</td><td>····················</td><td></td><td></td><td></td></tr>
<tr><td>C</td><td>8000</td><td>hCoV-19/USA/CA-UCSF-UC45/2020|EPI_ISL_450237|2020-</td><td>····················</td><td>····················</td><td>····················</td><td>····················</td><td>····················</td><td>····················</td><td></td><td></td><td></td></tr>
<tr><td>C</td><td>8001</td><td>hCoV-19/USA/CA-UCSF-UC46/2020|EPI_ISL_450238|2020-</td><td>····················</td><td>····················</td><td>····················</td><td>····················</td><td>····················</td><td>····················</td><td></td><td></td><td></td></tr>
<tr><td>C</td><td>8002</td><td>hCoV-19/USA/CA-UCSF-UC47/2020|EPI_ISL_450239|2020-</td><td>····················</td><td>·A··················</td><td>····················</td><td>····················</td><td>····················</td><td>····················</td><td></td><td></td><td></td></tr>
<tr><td>C</td><td>8003</td><td>hCoV-19/USA/CA-UCSF-UC48/2020|EPI_ISL_450240|2020-</td><td>····················</td><td>····················</td><td>····················</td><td>····················</td><td>····················</td><td>····················</td><td></td><td></td><td></td></tr>
<tr><td>C</td><td>8004</td><td>hCoV-19/Russia/StPetersburg-RII6251S/2020|EPI_ISL_</td><td>····················</td><td>····················</td><td>····················</td><td>····················</td><td>····················</td><td>····················</td><td></td><td></td><td></td></tr>
<tr><td>C</td><td>8005</td><td>hCoV-19/Russia/StPetersburg-RII6704V/2020|EPI_ISL_</td><td>····················</td><td>····················</td><td>····················</td><td>····················</td><td>····················</td><td>····················</td><td></td><td></td><td></td></tr>
<tr><td>C</td><td>8006</td><td>hCoV-19/Russia/StPetersburg-RII6790V/2020|EPI_ISL_</td><td>····················</td><td>····················</td><td>····················</td><td>····················</td><td>····················</td><td>····················</td><td></td><td></td><td></td></tr>
<tr><td>C</td><td>8007</td><td>hCoV-19/Russia/StPetersburg-RII6840S/2020|EPI_ISL_</td><td>····················</td><td>····················</td><td>····················</td><td>····················</td><td>····················</td><td>····················</td><td></td><td></td><td></td></tr>
<tr><td>C</td><td>8008</td><td>hCoV-19/Russia/StPetersburg-RII6846V/2020|EPI_ISL_</td><td>····················</td><td>····················</td><td>····················</td><td>····················</td><td>····················</td><td>····················</td><td></td><td></td><td></td></tr>
<tr><td>C</td><td>8009</td><td>hCoV-19/Russia/Kipen-RII6907V/2020|EPI_ISL_450246|</td><td>····················</td><td>····················</td><td>····················</td><td>····················</td><td>····················</td><td>····················</td><td></td><td></td><td></td></tr>
<tr><td>C</td><td>8010</td><td>hCoV-19/Russia/StPetersburg-RII7039V/2020|EPI_ISL_</td><td>····················</td><td>····················</td><td>····················</td><td>····················</td><td>····················</td><td>····················</td><td></td><td></td><td></td></tr>
<tr><td>C</td><td>8011</td><td>hCoV-19/Russia/StPetersburg-RII7318V/2020|EPI_ISL_</td><td>····················</td><td>····················</td><td>····················</td><td>····················</td><td>····················</td><td>····················</td><td></td><td></td><td></td></tr>
<tr><td>C</td><td>8012</td><td>hCoV-19/Russia/StPetersburg-RII7319V/2020|EPI_ISL_</td><td>····················</td><td>····················</td><td>····················</td><td>····················</td><td>····················</td><td>····················</td><td></td><td></td><td></td></tr>
<tr><td>C</td><td>8013</td><td>hCoV-19/Russia/StPetersburg-RII7321V/2020|EPI_ISL_</td><td>····················</td><td>····················</td><td>····················</td><td>····················</td><td>····················</td><td>····················</td><td></td><td></td><td></td></tr>
<tr><td>C</td><td>8014</td><td>hCoV-19/Russia/StPetersburg-RII7340V/2020|EPI_ISL_</td><td>····················</td><td>····················</td><td>····················</td><td>····················</td><td>····················</td><td>····················</td><td></td><td></td><td></td></tr>
<tr><td>C</td><td>8015</td><td>hCoV-19/Russia/StPetersburg-RII7464S/2020|EPI_ISL_</td><td>····················</td><td>····················</td><td>····················</td><td>····················</td><td>····················</td><td>····················</td><td></td><td></td><td></td></tr>
<tr><td>C</td><td>8016</td><td>hCoV-19/Russia/StPetersburg-RII7488S/2020|EPI_ISL_</td><td>····················</td><td>····················</td><td>····················</td><td>····················</td><td>····················</td><td>····················</td><td></td><td></td><td></td></tr>
<tr><td>C</td><td>8017</td><td>hCoV-19/Russia/StPetersburg-RII7501S/2020|EPI_ISL_</td><td>····················</td><td>····················</td><td>····················</td><td>····················</td><td>····················</td><td>····················</td><td></td><td></td><td></td></tr>
<tr><td>C</td><td>8018</td><td>hCoV-19/Russia/StPetersburg-RII7505S/2020|EPI_ISL_</td><td>····················</td><td>····················</td><td>····················</td><td>····················</td><td>····················</td><td>····················</td><td></td><td></td><td></td></tr>
<tr><td>C</td><td>8019</td><td>hCoV-19/Russia/StPetersburg-RII7520S/2020|EPI_ISL_</td><td>····················</td><td>····················</td><td>····················</td><td>····················</td><td>····················</td><td>····················</td><td></td><td></td><td></td></tr>
<tr><td>C</td><td>8020</td><td>hCoV-19/Russia/StPetersburg-RII7545S/2020|EPI_ISL_</td><td>····················</td><td>····················</td><td>····················</td><td>····················</td><td>····················</td><td>····················</td><td></td><td></td><td></td></tr>
<tr><td>C</td><td>8021</td><td>hCoV-19/Russia/StPetersburg-RII7553S/2020|EPI_ISL_</td><td>····················</td><td>····················</td><td>····················</td><td>····················</td><td>····················</td><td>····················</td><td></td><td></td><td></td></tr>
<tr><td>C</td><td>8022</td><td>hCoV-19/Russia/StPetersburg-RII7555S/2020|EPI_ISL_</td><td>····················</td><td>····················</td><td>····················</td><td>····················</td><td>····················</td><td>····················</td><td></td><td></td><td></td></tr>
</table>

C 8023 hCoV-19/Russia/StPetersburg-RII7557S/2020|EPI_ISL_ ···················· ···················· ···················· ···················· ···················· ····················
C 8024 hCoV-19/Russia/StPetersburg-RII7581S/2020|EPI_ISL_ ···················· ···················· ···················· ···················· ···················· ····················
C 8025 hCoV-19/Russia/StPetersburg-RII7594S/2020|EPI_ISL_ ···················· ···················· ···················· ···················· ···················· ····················
C 8026 hCoV-19/Russia/StPetersburg-RII7603S/2020|EPI_ISL_ ···················· ···················· ···················· ···················· ···················· ····················
C 8027 hCoV-19/Russia/StPetersburg-RII7656S/2020|EPI_ISL_ ···················· ···················· ···················· ···················· ···················· ····················
C 8028 hCoV-19/Russia/StPetersburg-RII7658S/2020|EPI_ISL_ ···················· ···················· ···················· ···················· ···················· ····················
C 8029 hCoV-19/Russia/StPetersburg-RII7880S/2020|EPI_ISL_ ···················· ···················· ···················· ···················· ···················· ····················
C 8030 hCoV-19/Russia/StPetersburg-RII7896S/2020|EPI_ISL_ ···················· ···················· ···················· ···················· ···················· ····················
C 8031 hCoV-19/Russia/StPetersburg-RII8275S/2020|EPI_ISL_ ···················· ···················· ···················· ···················· ···················· ····················
C 8032 hCoV-19/Russia/StPetersburg-RII8289S/2020|EPI_ISL_ ···················· ···················· ···················· ···················· ···················· ····················
C 8033 hCoV-19/Russia/StPetersburg-RII8298S/2020|EPI_ISL_ ···················· ···················· ···················· ···················· ···················· ····················
C 8034 hCoV-19/Russia/StPetersburg-RII8317S/2020|EPI_ISL_ ···················· ···················· ···················· ···················· ···················· ····················
C 8035 hCoV-19/Russia/StPetersburg-RII8319S/2020|EPI_ISL_ ···················· ···················· ···················· ···················· ···················· ····················
C 8036 hCoV-19/Russia/StPetersburg-RII8350S/2020|EPI_ISL_ ···················· ···················· ···················· ···················· ···················· ····················
C 8037 hCoV-19/Russia/StPetersburg-RII8371S/2020|EPI_ISL_ ···················· ···················· ···················· ···················· ···················· ····················
C 8038 hCoV-19/Russia/StPetersburg-RII8373S/2020|EPI_ISL_ ···················· ···················· ···················· ···················· ···················· ····················
C 8039 hCoV-19/Russia/StPetersburg-RII8380S/2020|EPI_ISL_ ···················· ···················· ···················· ···················· ···················· ····················
C 8040 hCoV-19/Russia/StPetersburg-RII8372S/2020|EPI_ISL_ ···················· ···················· ···················· ···················· ···················· ····················
C 8041 hCoV-19/Russia/StPetersburg-RII8388S/2020|EPI_ISL_ ···················· ···················· ···················· ···················· ···················· ····················
C 8042 hCoV-19/Russia/StPetersburg-RII8393S/2020|EPI_ISL_ ···················· ···················· ···················· ···················· ···················· ····················
C 8043 hCoV-19/Russia/StPetersburg-RII8397S/2020|EPI_ISL_ ···················· ···················· ···················· ···················· ···················· ····················
C 8044 hCoV-19/Russia/StPetersburg-RII8402S/2020|EPI_ISL_ ···················· ···················· ···················· ···················· ···A················ ····················
C 8045 hCoV-19/Russia/StPetersburg-RII8409S/2020|EPI_ISL_ ···················· ···················· ···················· ···················· ···················· ····················
C 8046 hCoV-19/Russia/StPetersburg-RII8417S/2020|EPI_ISL_ ···················· ···················· ···················· ···················· ···················· ····················
C 8047 hCoV-19/Russia/StPetersburg-RII8418S/2020|EPI_ISL_ ···················· ···················· ···················· ···················· ···················· ····················
C 8048 hCoV-19/Russia/StPetersburg-RII8425S/2020|EPI_ISL_ ···················· ···················· ···················· ···················· ···················· ····················
C 8049 hCoV-19/Russia/StPetersburg-RII8428S/2020|EPI_ISL_ ···················· ···················· ···················· ···················· ···················· ····················
C 8050 hCoV-19/Russia/StPetersburg-RII8429S/2020|EPI_ISL_ ···················· ···················· ···················· ···················· ···················· ····················
C 8051 hCoV-19/Russia/StPetersburg-RII8876S/2020|EPI_ISL_ ···················· ···················· ···················· ···················· ···················· ····················
C 8052 hCoV-19/Russia/StPetersburg-RII8890S/2020|EPI_ISL_ ···················· ···················· ···················· ···················· ···················· ····················
C 8053 hCoV-19/Russia/StPetersburg-RII8913S/2020|EPI_ISL_ ···················· ···················· ···················· ···················· ···················· ····················
C 8054 hCoV-19/Russia/StPetersburg-RII8934S/2020|EPI_ISL_ ···················· ···················· ···················· ···················· ···················· ····················
C 8055 hCoV-19/Russia/StPetersburg-RII8935S/2020|EPI_ISL_ ···················· ···················· ···················· ···················· ···················· ····················
C 8056 hCoV-19/Russia/StPetersburg-RII8955S/2020|EPI_ISL_ ···················· ···················· ···················· ···················· ···················· ····················
C 8057 hCoV-19/Poland/IHG_PAS_1_69/2020|EPI_ISL_450294|20 ···················· ···················· ···················· ···················· ···················· ····················
C 8058 hCoV-19/Poland/IHG_PAS_1_73/2020|EPI_ISL_450295|20 ···················· ···················· ···················· ···················· ···················· ····················
C 8059 hCoV-19/Australia/NSW277/2020|EPI_ISL_451541|2020- ···················· ···················· ···················· ···················· ···················· ····················
C 8060 hCoV-19/Australia/NSW278/2020|EPI_ISL_451542|2020- ···················· ···················· ···················· ···················· ···················· ····················
C 8061 hCoV-19/Australia/NSW279/2020|EPI_ISL_451543|2020- ···················· ···················· ···················· ···················· ···················· ····················
C 8062 hCoV-19/Australia/NSW280/2020|EPI_ISL_451544|2020- ···················· ···················· ···················· ···················· ···················· ····················
C 8063 hCoV-19/Australia/NSW281/2020|EPI_ISL_451545|2020- ···················· ···················· ···················· ···················· ···················· ····················
C 8064 hCoV-19/Australia/NSW282/2020|EPI_ISL_451546|2020- ···················· ···················· ···················· ···················· ···················· ····················
C 8065 hCoV-19/Australia/NSW283/2020|EPI_ISL_451547|2020- ···················· ···················· ···················· ···················· ···················· ····················
C 8066 hCoV-19/Australia/NSW284/2020|EPI_ISL_451548|2020- ···················· ···················· ···················· ···················· ···················· ····················
C 8067 hCoV-19/Australia/NSW285/2020|EPI_ISL_451549|2020- ···················· ···················· ···················· ···················· ···················· ····················
C 8068 hCoV-19/Australia/NSW286/2020|EPI_ISL_451550|2020- ···················· ···················· ···················· ···················· ···················· ····················
C 8069 hCoV-19/Australia/NSW287/2020|EPI_ISL_451551|2020- ···················· ···················· ···················· ···················· ···················· ····················
C 8070 hCoV-19/Australia/NSW288/2020|EPI_ISL_451552|2020- ···················· ···················· ···················· ···················· ···················· ····················
C 8071 hCoV-19/Australia/NSW289/2020|EPI_ISL_451553|2020- ···················· ···················· ···················· ···················· ···················· ····················
C 8072 hCoV-19/Australia/NSW290/2020|EPI_ISL_451554|2020- ···················· ···················· ···················· ···················· ···················· ····················
C 8073 hCoV-19/Australia/NSW291/2020|EPI_ISL_451555|2020- ···················· ···················· ···················· ···················· ···················· ····················
C 8074 hCoV-19/Australia/NSW292/2020|EPI_ISL_451556|2020- ···················· ···················· ···················· ···················· ···················· ····················
C 8075 hCoV-19/Australia/NSW293/2020|EPI_ISL_451557|2020- ···················· ···················· ···················· ···················· ···················· ····················
C 8076 hCoV-19/Australia/NSW294/2020|EPI_ISL_451558|2020- ···················· ···················· ···················· ···················· ···················· ····················
C 8077 hCoV-19/Australia/NSW295/2020|EPI_ISL_451559|2020- ···················· ···················· ···················· ···················· ···················· ····················
C 8078 hCoV-19/Australia/NSW296/2020|EPI_ISL_451560|2020- ···················· ···················· ···················· ···················· ···················· ····················
C 8079 hCoV-19/Australia/NSW297/2020|EPI_ISL_451561|2020- ···················· ···················· ···················· ···················· ···················· ····················
C 8080 hCoV-19/Australia/NSW298/2020|EPI_ISL_451562|2020- ···················· ···················· ···················· ···················· ···················· ····················
C 8081 hCoV-19/Australia/NSW299/2020|EPI_ISL_451563|2020- ···················· ···················· ···················· ···················· ···················· ····················
C 8082 hCoV-19/Australia/NSW300/2020|EPI_ISL_451564|2020- ···················· ···················· ···················· ···················· ···················· ····················
C 8083 hCoV-19/Australia/NSW301/2020|EPI_ISL_451565|2020- ···················· ···················· ···················· ···················· ···················· ····················
C 8084 hCoV-19/Australia/NSW302/2020|EPI_ISL_451566|2020- ···················· ···················· ···················· ···················· ···················· ····················
C 8085 hCoV-19/Australia/NSW303/2020|EPI_ISL_451567|2020- ···················· ···················· ···················· ···················· ···················· ····················
C 8086 hCoV-19/Australia/NSW304/2020|EPI_ISL_451568|2020- ···················· ···················· ···················· ···················· ···················· ····················
C 8087 hCoV-19/Australia/NSW305/2020|EPI_ISL_451569|2020- ···················· ···················· ···················· ···················· ···················· ····················
C 8088 hCoV-19/Australia/NSW306/2020|EPI_ISL_451570|2020- ···················· ···················· ···················· ···················· ···················· ····················
C 8089 hCoV-19/Australia/NSW307/2020|EPI_ISL_451571|2020- ···················· ···················· ···················· ···················· ···················· ····················
C 8090 hCoV-19/Australia/NSW308/2020|EPI_ISL_451572|2020- ···················· ···················· ···················· ···················· ···················· ····················
C 8091 hCoV-19/Australia/NSW309/2020|EPI_ISL_451573|2020- ···················· ···················· ···················· ···················· ···················· ····················
C 8092 hCoV-19/Australia/NSW310/2020|EPI_ISL_451574|2020- ···················· ···················· ···················· ···················· ···················· ····················
C 8093 hCoV-19/Australia/NSW311/2020|EPI_ISL_451575|2020- ···················· ···················· ···················· ···················· ···················· ····················
C 8094 hCoV-19/Australia/NSW312/2020|EPI_ISL_451576|2020- ···················· ···················· ···················· ···················· ···················· ····················
C 8095 hCoV-19/Australia/NSW314/2020|EPI_ISL_451577|2020- ···················· ···················· ···················· ···················· ···················· ····················
C 8096 hCoV-19/Australia/NSW315/2020|EPI_ISL_451578|2020- ···················· ···················· ···················· ···················· ···················· ····················
C 8097 hCoV-19/Australia/NSW316/2020|EPI_ISL_451579|2020- ···················· ···················· ···················· ···················· ···················· ····················
C 8098 hCoV-19/Australia/NSW317/2020|EPI_ISL_451580|2020- ···················· ···················· ···················· ···················· ···················· ····················
C 8099 hCoV-19/Australia/NSW318/2020|EPI_ISL_451581|2020- ···················· ···················· ···················· ···················· ···················· ····················
C 8100 hCoV-19/Australia/NSW319/2020|EPI_ISL_451582|2020- ···················· ···················· ···················· ···················· ···················· ····················

| | | | | | | | | |
|---|---|---|---|---|---|---|---|---|
| C | 8101 | hCoV-19/Australia/NSW321/2020\|EPI_ISL_451583\|2020- | .................... | .................... | .................... | .................... | .................... | .................... |
| C | 8102 | hCoV-19/Australia/NSW322/2020\|EPI_ISL_451584\|2020- | .................... | .................... | .................... | .................... | .................... | .................... |
| C | 8103 | hCoV-19/Australia/NSW324/2020\|EPI_ISL_451586\|2020- | .................... | .................... | .................... | .................... | .................... | .................... |
| C | 8104 | hCoV-19/Australia/NSW325/2020\|EPI_ISL_451587\|2020- | .................... | .................... | .................... | .................... | .................... | .................... |
| C | 8105 | hCoV-19/Australia/NSW326/2020\|EPI_ISL_451588\|2020- | .................... | .................... | .................... | .................... | .................... | .................... |
| C | 8106 | hCoV-19/Australia/NSW327/2020\|EPI_ISL_451589\|2020- | .................... | .................... | .................... | .................... | .................... | .................... |
| C | 8107 | hCoV-19/Australia/NSW328/2020\|EPI_ISL_451590\|2020- | .................... | .................... | .................... | .................... | .................... | .................... |
| C | 8108 | hCoV-19/Australia/NSW329/2020\|EPI_ISL_451591\|2020- | .................... | .................... | .................... | .................... | .................... | .................... |
| C | 8109 | hCoV-19/Australia/NSW330/2020\|EPI_ISL_451592\|2020- | .................... | .................... | .................... | .................... | .................... | .................... |
| C | 8110 | hCoV-19/Australia/NSW331/2020\|EPI_ISL_451593\|2020- | .................... | .................... | .................... | .................... | .................... | .................... |
| C | 8111 | hCoV-19/Australia/NSW332/2020\|EPI_ISL_451594\|2020- | .................... | .................... | .................... | .................... | .................... | .................... |
| C | 8112 | hCoV-19/Australia/NSW333/2020\|EPI_ISL_451595\|2020- | .................... | .................... | .................... | .................... | .................... | .................... |
| C | 8113 | hCoV-19/Australia/NSW334/2020\|EPI_ISL_451596\|2020- | .................... | .................... | .................... | .................... | .................... | .................... |
| C | 8114 | hCoV-19/Australia/NSW335/2020\|EPI_ISL_451597\|2020- | .................... | .................... | .................... | .................... | .................... | .................... |
| C | 8115 | hCoV-19/Australia/NSW336/2020\|EPI_ISL_451598\|2020- | .................... | .................... | .................... | .................... | .................... | .................... |
| C | 8116 | hCoV-19/Australia/NSW337/2020\|EPI_ISL_451599\|2020- | .................... | .................... | .................... | .................... | .................... | .................... |
| C | 8117 | hCoV-19/USA/VA-DCLS-0246/2020\|EPI_ISL_452820\|2020- | .................... | .................... | .................... | .................... | .................... | .................... |
| C | 8118 | hCoV-19/USA/VA-DCLS-0247/2020\|EPI_ISL_452821\|2020- | .................... | .................... | .................... | .................... | .................... | .................... |
| C | 8119 | hCoV-19/USA/VA-DCLS-0248/2020\|EPI_ISL_452822\|2020- | .................... | .................... | .................... | .................... | .................... | .................... |
| C | 8120 | hCoV-19/USA/VA-DCLS-0249/2020\|EPI_ISL_452823\|2020- | .................... | .................... | .................... | .................... | .................... | .................... |
| C | 8121 | hCoV-19/USA/VA-DCLS-0251/2020\|EPI_ISL_452825\|2020- | .................... | .................... | .................... | .................... | .................... | .................... |
| C | 8122 | hCoV-19/USA/VA-DCLS-0252/2020\|EPI_ISL_452826\|2020- | .................... | .................... | .................... | .................... | .................... | .................... |
| C | 8123 | hCoV-19/USA/VA-DCLS-0253/2020\|EPI_ISL_452827\|2020- | .................... | .................... | .................... | .................... | .................... | .................... |
| C | 8124 | hCoV-19/USA/VA-DCLS-0254/2020\|EPI_ISL_452828\|2020- | .................... | .................... | .................... | .................... | .................... | .................... |
| C | 8125 | hCoV-19/USA/VA-DCLS-0255/2020\|EPI_ISL_452829\|2020- | .................... | .................... | .................... | .................... | .................... | .................... |
| C | 8126 | hCoV-19/England/CAMB-1AFC95/2020\|EPI_ISL_452872\|20 | .................... | .................... | .................... | .................... | .................... | .................... |
| C | 8127 | hCoV-19/England/CAMB-1AFD1A/2020\|EPI_ISL_452876\|20 | .................... | .................... | .................... | .................... | .................... | .................... |
| C | 8128 | hCoV-19/England/CAMB-1AFD38/2020\|EPI_ISL_452877\|20 | .................... | .................... | .................... | .................... | .................... | .................... |
| C | 8129 | hCoV-19/England/CAMB-1AFD56/2020\|EPI_ISL_452878\|20 | .................... | .................... | .................... | .................... | .................... | .................... |
| C | 8130 | hCoV-19/England/CAMB-1AFD74/2020\|EPI_ISL_452879\|20 | .................... | .................... | .................... | .................... | .................... | .................... |
| C | 8131 | hCoV-19/England/CAMB-1B0138/2020\|EPI_ISL_452880\|20 | .................... | .................... | .................... | .................... | .................... | .................... |
| C | 8132 | hCoV-19/England/CAMB-1B0147/2020\|EPI_ISL_452881\|20 | .................... | .................... | .................... | .................... | .................... | .................... |
| C | 8133 | hCoV-19/England/CAMB-1B0156/2020\|EPI_ISL_452882\|20 | .................... | .................... | .................... | .................... | .................... | .................... |
| C | 8134 | hCoV-19/England/CAMB-1B0183/2020\|EPI_ISL_452884\|20 | .................... | .................... | .................... | .................... | .................... | .................... |
| C | 8135 | hCoV-19/England/CAMB-1B04D5/2020\|EPI_ISL_452885\|20 | .................... | .................... | .................... | .................... | .................... | .................... |
| C | 8136 | hCoV-19/England/CAMB-1B04E4/2020\|EPI_ISL_452886\|20 | .................... | .................... | .................... | .................... | .................... | .................... |
| C | 8137 | hCoV-19/England/CAMB-1B052D/2020\|EPI_ISL_452888\|20 | .................... | .................... | .................... | .................... | .................... | .................... |
| C | 8138 | hCoV-19/England/CAMB-1B053C/2020\|EPI_ISL_452889\|20 | .................... | .................... | .................... | .................... | .................... | .................... |
| C | 8139 | hCoV-19/England/CAMB-1B054B/2020\|EPI_ISL_452890\|20 | .................... | .................... | .................... | .................... | .................... | .................... |
| C | 8140 | hCoV-19/England/CAMB-1B055A/2020\|EPI_ISL_452891\|20 | .................... | .................... | .................... | .................... | .................... | .................... |
| C | 8141 | hCoV-19/England/CAMB-1B0569/2020\|EPI_ISL_452892\|20 | .................... | .................... | .................... | .................... | .................... | .................... |
| C | 8142 | hCoV-19/England/CAMB-1B0578/2020\|EPI_ISL_452893\|20 | .................... | .................... | .................... | .................... | .................... | .................... |
| C | 8143 | hCoV-19/England/CAMB-1B0596/2020\|EPI_ISL_452894\|20 | .................... | .................... | .................... | .................... | .................... | .................... |
| C | 8144 | hCoV-19/England/CAMB-1B05A5/2020\|EPI_ISL_452895\|20 | .................... | .................... | .................... | .................... | .................... | .................... |
| C | 8146 | hCoV-19/England/CAMB-1B05C3/2020\|EPI_ISL_452897\|20 | .................... | .................... | .................... | .................... | .................... | .................... |
| C | 8147 | hCoV-19/England/CAMB-1B05E1/2020\|EPI_ISL_452898\|20 | .................... | .................... | .................... | .................... | .................... | .................... |
| C | 8148 | hCoV-19/Sweden/20-06761/2020\|EPI_ISL_455100\|2020-0 | .................... | .................... | .................... | .................... | .................... | .................... |
| C | 8149 | hCoV-19/Sweden/20-07377/2020\|EPI_ISL_455101\|2020-0 | .................... | .................... | .................... | .................... | .................... | .................... |
| C | 8150 | hCoV-19/Sweden/20-07896/2020\|EPI_ISL_455102\|2020-0 | .................... | .................... | .................... | .................... | .................... | .................... |
| C | 8151 | hCoV-19/Sweden/20-14013/2020\|EPI_ISL_455103\|2020-0 | .................... | .................... | .................... | .................... | .................... | .................... |
| C | 8152 | hCoV-19/Sweden/20-51264/2020\|EPI_ISL_455104\|2020-0 | .................... | .................... | .................... | .................... | .................... | .................... |
| C | 8153 | hCoV-19/New Zealand/20VR3045/2020\|EPI_ISL_456402\|2 | .................... | .................... | .................... | .................... | .................... | .................... |
| C | 8154 | hCoV-19/New Zealand/20VR3064/2020\|EPI_ISL_456403\|2 | .................... | .................... | .................... | .................... | .................... | .................... |
| C | 8165 | hCoV-19/Estonia/ChVir4285/2020\|EPI_ISL_457723\|2020 | .................... | .................... | .................... | .................... | .................... | .................... |
| C | 8166 | hCoV-19/Italy/AVA-AMVRC-4875/2020\|EPI_ISL_457724\|2 | .................... | .................... | .................... | .................... | .................... | .................... |
| C | 8167 | hCoV-19/Estonia/ChVir4286/2020\|EPI_ISL_457725\|2020 | .................... | .................... | .................... | .................... | .................... | .................... |
| C | 8168 | hCoV-19/Taiwan/TSGH-35/2020\|EPI_ISL_457726\|2020-02 | .................... | .................... | .................... | .................... | .................... | .................... |
| C | 8169 | hCoV-19/Estonia/ChVir4287/2020\|EPI_ISL_457727\|2020 | .................... | .................... | .................... | .................... | .................... | .................... |
| C | 8170 | hCoV-19/Italy/MOL-AMVRC-2389/2020\|EPI_ISL_457728\|2 | .................... | .................... | .................... | .................... | .................... | .................... |
| C | 8171 | hCoV-19/Estonia/ChVir4292/2020\|EPI_ISL_457729\|2020 | .................... | .................... | .................... | .................... | .................... | .................... |
| C | 8172 | hCoV-19/Taiwan/TSGH-36/2020\|EPI_ISL_457730\|2020-02 | .................... | .................... | .................... | .................... | .................... | .................... |
| C | 8173 | hCoV-19/Italy/AVA-AMVRC-4870/2020\|EPI_ISL_457732\|2 | .................... | .................... | .................... | .................... | .................... | .................... |
| C | 8174 | hCoV-19/Taiwan/TSGH-37/2020\|EPI_ISL_457733\|2020-02 | .................... | .................... | .................... | .................... | .................... | .................... |
| C | 8175 | hCoV-19/Estonia/ChVir4299/2020\|EPI_ISL_457735\|2020 | .................... | .................... | .................... | .................... | .................... | .................... |
| C | 8176 | hCoV-19/Italy/BAS-UniMI-5062/2020\|EPI_ISL_457736\|2 | .................... | .................... | .................... | .................... | .................... | .................... |
| C | 8177 | hCoV-19/Estonia/ChVir4301/2020\|EPI_ISL_457737\|2020 | .................... | .................... | .................... | .................... | .................... | .................... |
| C | 8178 | hCoV-19/Estonia/ChVir4313/2020\|EPI_ISL_457740\|2020 | .................... | .................... | .................... | .................... | .................... | .................... |
| C | 8179 | hCoV-19/Italy/ABR-IZSGC-484/2020\|EPI_ISL_457749\|20 | .................... | .................... | .................... | .................... | .................... | .................... |
| C | 8180 | hCoV-19/Germany/MV-Cento-COV7/2020\|EPI_ISL_457750\| | .................... | .................... | .................... | .................... | .................... | .................... |
| C | 8181 | hCoV-19/USA/MD-HP00029/2020\|EPI_ISL_457752\|2020-03 | .................... | .................... | .................... | .................... | .................... | .................... |
| C | 8182 | hCoV-19/USA/DC-HP00035/2020\|EPI_ISL_457753\|2020-03 | .................... | .................... | .................... | .................... | .................... | .................... |
| C | 8183 | hCoV-19/USA/MD-HP00039/2020\|EPI_ISL_457754\|2020-03 | .................... | .................... | .................... | .................... | .................... | .................... |
| C | 8188 | hCoV-19/USA/DC-HP00096/2020\|EPI_ISL_457760\|2020-03 | .................... | .................... | .................... | .................... | .................... | .................... |
| C | 8192 | hCoV-19/USA/MD-HP00110/2020\|EPI_ISL_457766\|2020-03 | .................... | .................... | .................... | .................... | .................... | .................... |
| C | 8193 | hCoV-19/Scotland/CVR3170/2020\|EPI_ISL_449200\|2020- | .................... | .................... | .................... | .................... | .................... | .................... |
| C | 8194 | hCoV-19/Scotland/CVR3171/2020\|EPI_ISL_449201\|2020- | .................... | .................... | .................... | .................... | .................... | .................... |
| C | 8195 | hCoV-19/Scotland/CVR3175/2020\|EPI_ISL_449203\|2020- | .................... | .................... | .................... | .................... | .................... | .................... |
| C | 8196 | hCoV-19/Scotland/CVR3176/2020\|EPI_ISL_449204\|2020- | .................... | .................... | .................... | .................... | .................... | .................... |

| | | | | | | | | |
|---|---|---|---|---|---|---|---|---|
| C | 8197 hCoV-19/Scotland/CVR3178/2020\|EPI_ISL_449206\|2020- | ···················· | ···················· | ···················· | ···················· | ···················· | ···················· | |
| C | 8198 hCoV-19/Scotland/CVR3182/2020\|EPI_ISL_449207\|2020- | ···················· | ···················· | ···················· | ···················· | ···················· | ···················· | |
| C | 8199 hCoV-19/Scotland/CVR3184/2020\|EPI_ISL_449208\|2020- | ···················· | ···················· | ···················· | ···················· | ···················· | ···················· | |
| C | 8200 hCoV-19/Scotland/CVR3185/2020\|EPI_ISL_449209\|2020- | ···················· | ···················· | ···················· | ···················· | ···················· | ···················· | |
| C | 8201 hCoV-19/Scotland/CVR3186/2020\|EPI_ISL_449210\|2020- | ···················· | ···················· | ···················· | ···················· | ···················· | ···················· | |
| C | 8202 hCoV-19/Scotland/CVR3189/2020\|EPI_ISL_449211\|2020- | ···················· | ·A·················· | ···················· | ···················· | ···················· | ···················· | |
| C | 8203 hCoV-19/Scotland/CVR3191/2020\|EPI_ISL_449212\|2020- | ···················· | ··············C····· | ···················· | ···················· | ···················· | ···················· | |
| C | 8204 hCoV-19/Scotland/CVR3193/2020\|EPI_ISL_449213\|2020- | ···················· | ···················· | ···················· | ···················· | ···················· | ···················· | |
| C | 8205 hCoV-19/Scotland/CVR3194/2020\|EPI_ISL_449214\|2020- | ···················· | ···················· | ···················· | ···················· | ···················· | ···················· | |
| C | 8206 hCoV-19/Scotland/CVR3195/2020\|EPI_ISL_449215\|2020- | ···················· | ···················· | ···················· | ···················· | ···················· | ···················· | |
| C | 8207 hCoV-19/Scotland/CVR3196/2020\|EPI_ISL_449216\|2020- | ···················· | ···················A | ···················· | ···················· | ···················· | ···················· | 1 |
| C | 8208 hCoV-19/Scotland/CVR3197/2020\|EPI_ISL_449217\|2020- | ···················· | ···················· | ···················· | ···················· | ···················· | ···················· | |
| C | 8209 hCoV-19/Scotland/CVR3198/2020\|EPI_ISL_449218\|2020- | ···················· | ···················· | ···················· | ···················· | ···················· | ···················· | |
| C | 8210 hCoV-19/Scotland/CVR3199/2020\|EPI_ISL_449219\|2020- | ···················· | ···················· | ···················· | ···················· | ···················· | ···················· | |
| C | 8211 hCoV-19/Scotland/CVR3200/2020\|EPI_ISL_449220\|2020- | ···················· | ···················A | ···················· | ···················· | ···················· | ···················· | 1 |
| C | 8212 hCoV-19/Scotland/CVR3202/2020\|EPI_ISL_449221\|2020- | ···················· | ···················· | ···················· | ···················· | ···················· | ···················· | |
| C | 8213 hCoV-19/Scotland/CVR3203/2020\|EPI_ISL_449222\|2020- | ···················· | ···················· | ···················· | ···················· | ···················· | ···················· | |
| C | 8214 hCoV-19/Scotland/CVR3204/2020\|EPI_ISL_449223\|2020- | ···················· | ···················A | ···················· | ···················· | ···················· | ···················· | 1 |
| C | 8215 hCoV-19/Scotland/CVR3205/2020\|EPI_ISL_449224\|2020- | ···················· | ···················· | ···················· | ···················· | ···················· | ···················· | |
| C | 8216 hCoV-19/Scotland/CVR3206/2020\|EPI_ISL_449225\|2020- | ···················· | ···················· | ···················· | ···················· | ···················· | ···················· | |
| C | 8217 hCoV-19/Scotland/CVR3207/2020\|EPI_ISL_449226\|2020- | ···················· | ···················· | ···················· | ···················· | ···················· | ···················· | |
| C | 8218 hCoV-19/Scotland/CVR3208/2020\|EPI_ISL_449227\|2020- | ···················· | ···················· | ···················· | ···················· | ···················· | ···················· | |
| C | 8219 hCoV-19/Scotland/CVR3209/2020\|EPI_ISL_449228\|2020- | ···················· | ···················· | ···················· | ···················· | ···················· | ···················· | |
| C | 8220 hCoV-19/Scotland/CVR3210/2020\|EPI_ISL_449229\|2020- | ···················· | ···················· | ···················· | ···················· | ···················· | ···················· | |
| C | 8221 hCoV-19/Australia/NSW234/2020\|EPI_ISL_451500\|2020- | ···················· | ···················· | ···················· | ···················· | ···················· | ···················· | |
| C | 8222 hCoV-19/Australia/NSW235/2020\|EPI_ISL_451501\|2020- | ···················· | ···················· | ···················· | ···················· | ···················· | ···················· | |
| C | 8223 hCoV-19/Australia/NSW236/2020\|EPI_ISL_451502\|2020- | ···················· | ···················· | ···················· | ···················· | ···················· | ···················· | |
| C | 8224 hCoV-19/Australia/NSW237/2020\|EPI_ISL_451503\|2020- | ···················· | ···················· | ···················· | ···················· | ···················· | ···················· | |
| C | 8225 hCoV-19/Australia/NSW238/2020\|EPI_ISL_451504\|2020- | ···················· | ···················· | ···················· | ···················· | ···················· | ···················· | |
| C | 8226 hCoV-19/Australia/NSW239/2020\|EPI_ISL_451505\|2020- | ···················· | ···················· | ···················· | ···················· | ···················· | ···················· | |
| C | 8227 hCoV-19/Australia/NSW240/2020\|EPI_ISL_451506\|2020- | ···················· | ···················· | ···················· | ···················· | ···················· | ···················· | |
| C | 8228 hCoV-19/Australia/NSW241/2020\|EPI_ISL_451507\|2020- | ···················· | ···················· | ···················· | ···················· | ···················· | ···················· | |
| C | 8229 hCoV-19/Australia/NSW242/2020\|EPI_ISL_451508\|2020- | ···················· | ···················· | ···················· | ···················· | ···················· | ···················· | |
| C | 8230 hCoV-19/Australia/NSW243/2020\|EPI_ISL_451509\|2020- | ···················· | ···················· | ···················· | ···················· | ···················· | ···················· | |
| C | 8231 hCoV-19/Australia/NSW245/2020\|EPI_ISL_451510\|2020- | ···················· | ···················· | ···················· | ···················· | ···················· | ···················· | |
| C | 8232 hCoV-19/Australia/NSW246/2020\|EPI_ISL_451511\|2020- | ···················· | ···················· | ···················· | ···················· | ···················· | ···················· | |
| C | 8233 hCoV-19/Australia/NSW247/2020\|EPI_ISL_451512\|2020- | ···················· | ···················· | ···················· | ···················· | ···················· | ···················· | |
| C | 8234 hCoV-19/Australia/NSW248/2020\|EPI_ISL_451513\|2020- | ···················· | ···················· | ···················· | ···················· | ···················· | ···················· | |
| C | 8235 hCoV-19/Australia/NSW249/2020\|EPI_ISL_451514\|2020- | ···················· | ···················· | ···················· | ···················· | ···················· | ···················· | |
| C | 8236 hCoV-19/Australia/NSW250/2020\|EPI_ISL_451515\|2020- | ···················· | ···················· | ···················· | ···················· | ···················· | ···················· | |
| C | 8237 hCoV-19/Australia/NSW251/2020\|EPI_ISL_451516\|2020- | ···················· | ···················· | ···················· | ···················· | ···················· | ···················· | |
| C | 8238 hCoV-19/Australia/NSW252/2020\|EPI_ISL_451517\|2020- | ···················· | ···················· | ···················· | ···················· | ···················· | ···················· | |
| C | 8239 hCoV-19/Australia/NSW253/2020\|EPI_ISL_451518\|2020- | ···················· | ···················· | ···················· | ···················· | ···················· | ···················· | |
| C | 8240 hCoV-19/Australia/NSW254/2020\|EPI_ISL_451519\|2020- | ···················· | ···················· | ···················· | ···················· | ···················· | ···················· | |
| C | 8241 hCoV-19/Australia/NSW256/2020\|EPI_ISL_451521\|2020- | ···················· | ···················· | ···················· | ···················· | ···················· | ···················· | |
| C | 8242 hCoV-19/Australia/NSW257/2020\|EPI_ISL_451522\|2020- | ···················· | ···················· | ···················· | ···················· | ···················· | ···················· | |
| C | 8243 hCoV-19/Australia/NSW258/2020\|EPI_ISL_451523\|2020- | ···················· | ···················· | ···················· | ···················· | ···················· | ···················· | |
| C | 8244 hCoV-19/Australia/NSW259/2020\|EPI_ISL_451524\|2020- | ···················· | ···················· | ···················· | ···················· | ···················· | ···················· | |
| C | 8245 hCoV-19/Australia/NSW260/2020\|EPI_ISL_451525\|2020- | ···················· | ···················· | ···················· | ···················· | ···················· | ···················· | |
| C | 8246 hCoV-19/Australia/NSW261/2020\|EPI_ISL_451526\|2020- | ···················· | ···················· | ···················· | ···················· | ···················· | ···················· | |
| C | 8247 hCoV-19/Australia/NSW262/2020\|EPI_ISL_451527\|2020- | ···················· | ···················· | ···················· | ···················· | ···················· | ···················· | |
| C | 8248 hCoV-19/Australia/NSW263/2020\|EPI_ISL_451528\|2020- | ···················· | ···················· | ···················· | ···················· | ···················· | ···················· | |
| C | 8249 hCoV-19/Australia/NSW264/2020\|EPI_ISL_451529\|2020- | ···················· | ···················· | ···················· | ···················· | ···················· | ···················· | |
| C | 8250 hCoV-19/Australia/NSW265/2020\|EPI_ISL_451530\|2020- | ···················· | ···················· | ···················· | ···················· | ···················· | ···················· | |
| C | 8251 hCoV-19/Australia/NSW266/2020\|EPI_ISL_451531\|2020- | ···················· | ···················· | ···················· | ···················· | ···················· | ···················· | |
| C | 8252 hCoV-19/Australia/NSW267/2020\|EPI_ISL_451532\|2020- | ···················· | ···················· | ···················· | ···················· | ···················· | ···················· | |
| C | 8253 hCoV-19/Australia/NSW268/2020\|EPI_ISL_451533\|2020- | ···················· | ···················· | ···················· | ···················· | ···················· | ···················· | |
| C | 8254 hCoV-19/Australia/NSW269/2020\|EPI_ISL_451534\|2020- | ···················· | ···················· | ···················· | ···················· | ···················· | ···················· | |
| C | 8255 hCoV-19/Australia/NSW270/2020\|EPI_ISL_451535\|2020- | ···················· | ···················· | ···················· | ···················· | ···················· | ···················· | |
| C | 8256 hCoV-19/Australia/NSW271/2020\|EPI_ISL_451536\|2020- | ···················· | ···················· | ···················· | ···················· | ···················· | ···················· | |
| C | 8257 hCoV-19/Australia/NSW272/2020\|EPI_ISL_451537\|2020- | ···················· | ···················· | ···················· | ···················· | ···················· | ···················· | |
| C | 8258 hCoV-19/Australia/NSW273/2020\|EPI_ISL_451538\|2020- | ···················· | ···················· | ···················· | ···················· | ···················· | ···················· | |
| C | 8259 hCoV-19/Australia/NSW275/2020\|EPI_ISL_451539\|2020- | ···················· | ···················· | ···················· | ···················· | ···················· | ···················· | |
| C | 8260 hCoV-19/Australia/NSW276/2020\|EPI_ISL_451540\|2020- | ···················· | ···················· | ···················· | ···················· | ···················· | ···················· | |
| C | 8261 hCoV-19/USA/VA-DCLS-0223/2020\|EPI_ISL_452800\|2020- | ···················· | ···················· | ···················· | ···················· | ···················· | ···················· | |
| C | 8262 hCoV-19/USA/VA-DCLS-0224/2020\|EPI_ISL_452801\|2020- | ···················· | ···················· | ···················· | ···················· | ···················· | ···················· | |
| C | 8263 hCoV-19/USA/VA-DCLS-0225/2020\|EPI_ISL_452802\|2020- | ···················· | ···················· | ···················· | ···················· | ···················· | ···················· | |
| C | 8264 hCoV-19/USA/VA-DCLS-0226/2020\|EPI_ISL_452803\|2020- | ···················· | ···················· | ···················· | ···················· | ···················· | ···················· | |
| C | 8265 hCoV-19/USA/VA-DCLS-0227/2020\|EPI_ISL_452804\|2020- | ···················· | ···················· | ···················· | ···················· | ···················· | ···················· | |
| C | 8266 hCoV-19/USA/VA-DCLS-0228/2020\|EPI_ISL_452805\|2020- | ···················· | ···················· | ···················· | ···················· | ···················· | ···················· | |
| C | 8267 hCoV-19/USA/VA-DCLS-0229/2020\|EPI_ISL_452806\|2020- | ···················· | ···················· | ···················· | ···················· | ···················· | ···················· | |
| C | 8268 hCoV-19/USA/VA-DCLS-0230/2020\|EPI_ISL_452807\|2020- | ···················· | ···················· | ···················· | ···················· | ···················· | ···················· | |
| C | 8269 hCoV-19/USA/VA-DCLS-0231/2020\|EPI_ISL_452808\|2020- | ···················· | ···················· | ···················· | ···················· | ···················· | ···················· | |
| C | 8270 hCoV-19/USA/VA-DCLS-0232/2020\|EPI_ISL_452809\|2020- | ···················· | ···················· | ···················· | ···················· | ···················· | ···················· | |
| C | 8271 hCoV-19/USA/VA-DCLS-0233/2020\|EPI_ISL_452810\|2020- | ···················· | ···················· | ···················· | ···················· | ···················· | ···················· | |
| C | 8272 hCoV-19/USA/VA-DCLS-0236/2020\|EPI_ISL_452811\|2020- | ···················· | ···················· | ···················· | ···················· | ···················· | ···················· | |
| C | 8273 hCoV-19/USA/VA-DCLS-0237/2020\|EPI_ISL_452812\|2020- | ···················· | ···················· | ···················· | ···················· | ···················· | ···················· | |
| C | 8274 hCoV-19/USA/VA-DCLS-0238/2020\|EPI_ISL_452813\|2020- | ···················· | ···················· | ···················· | ···················· | ···················· | ···················· | |

| | | | | | | | | |
|---|---|---|---|---|---|---|---|---|
| C | 8275 | hCoV-19/USA/VA-DCLS-0239/2020\|EPI_ISL_452814\|2020- | .................... | .................... | ..................... | .................... | .................... | .................... |
| C | 8276 | hCoV-19/USA/VA-DCLS-0240/2020\|EPI_ISL_452815\|2020- | .................... | .................... | ..................... | .................... | .................... | .................... |
| C | 8277 | hCoV-19/USA/VA-DCLS-0241/2020\|EPI_ISL_452816\|2020- | .................... | .................... | ..................... | .................... | .................... | .................... |
| C | 8278 | hCoV-19/USA/VA-DCLS-0243/2020\|EPI_ISL_452817\|2020- | .................... | .................... | ..................... | .................... | .................... | .................... |
| C | 8279 | hCoV-19/USA/VA-DCLS-0244/2020\|EPI_ISL_452818\|2020- | .................... | .................... | ..................... | .................... | .................... | .................... |
| C | 8280 | hCoV-19/USA/VA-DCLS-0245/2020\|EPI_ISL_452819\|2020- | .................... | .................... | ..................... | .................... | .................... | .................... |
| C | 8281 | hCoV-19/USA/VA-DCLS-0250/2020\|EPI_ISL_452824\|2020- | .................... | .................... | ..................... | .................... | .................... | .................... |
| C | 8282 | hCoV-19/USA/VA-DCLS-0257/2020\|EPI_ISL_452830\|2020- | .................... | .................... | ..................... | .................... | .................... | .................... |
| C | 8283 | hCoV-19/USA/VA-DCLS-0258/2020\|EPI_ISL_452831\|2020- | .................... | .................... | ..................... | .................... | .................... | .................... |
| C | 8284 | hCoV-19/USA/VA-DCLS-0259/2020\|EPI_ISL_452832\|2020- | .................... | .................... | ..................... | .................... | .................... | .................... |
| C | 8285 | hCoV-19/USA/VA-DCLS-0260/2020\|EPI_ISL_452833\|2020- | .................... | .................... | ..................... | .................... | .................... | .................... |
| C | 8286 | hCoV-19/USA/VA-DCLS-0261/2020\|EPI_ISL_452834\|2020- | .................... | .................... | ..................... | .................... | .................... | .................... |
| C | 8287 | hCoV-19/USA/VA-DCLS-0262/2020\|EPI_ISL_452835\|2020- | .................... | .................... | ..................... | .................... | .................... | .................... |
| C | 8288 | hCoV-19/USA/VA-DCLS-0263/2020\|EPI_ISL_452836\|2020- | .................... | .................... | ..................... | .................... | .................... | .................... |
| C | 8289 | hCoV-19/USA/VA-DCLS-0264/2020\|EPI_ISL_452837\|2020- | .................... | .................... | ..................... | .................... | .................... | .................... |
| C | 8290 | hCoV-19/USA/VA-DCLS-0265/2020\|EPI_ISL_452838\|2020- | .................... | .................... | ..................... | .................... | .................... | .................... |
| C | 8291 | hCoV-19/USA/VA-DCLS-0266/2020\|EPI_ISL_452839\|2020- | .................... | .................... | ..................... | .................... | .................... | .................... |
| C | 8292 | hCoV-19/USA/VA-DCLS-0271/2020\|EPI_ISL_452840\|2020- | .................... | .................... | ..................... | .................... | .................... | .................... |
| C | 8293 | hCoV-19/USA/VA-DCLS-0272/2020\|EPI_ISL_452841\|2020- | .................... | .................... | ..................... | .................... | .................... | .................... |
| C | 8294 | hCoV-19/USA/VA-DCLS-0273/2020\|EPI_ISL_452842\|2020- | .................... | .................... | ..................... | .................... | .................... | .................... |
| C | 8295 | hCoV-19/USA/VA-DCLS-0274/2020\|EPI_ISL_452843\|2020- | .................... | .................... | ..................... | .................... | .................... | .................... |
| C | 8296 | hCoV-19/USA/VA-DCLS-0275/2020\|EPI_ISL_452844\|2020- | .................... | .................... | ..................... | .................... | .................... | .................... |
| C | 8297 | hCoV-19/USA/VA-DCLS-0276/2020\|EPI_ISL_452845\|2020- | .................... | .................... | ..................... | .................... | .................... | .................... |
| C | 8298 | hCoV-19/USA/VA-DCLS-0277/2020\|EPI_ISL_452846\|2020- | .................... | .................... | ..................... | .................... | .................... | .................... |
| C | 8299 | hCoV-19/USA/VA-DCLS-0278/2020\|EPI_ISL_452847\|2020- | .................... | .................... | ..................... | .................... | .................... | .................... |
| C | 8300 | hCoV-19/USA/VA-DCLS-0279/2020\|EPI_ISL_452848\|2020- | .................... | .................... | ..................... | .................... | .................... | .................... |
| C | 8301 | hCoV-19/USA/VA-DCLS-0280/2020\|EPI_ISL_452849\|2020- | .................... | .................... | ..................... | .................... | .................... | .................... |
| C | 8302 | hCoV-19/USA/VA-DCLS-0281/2020\|EPI_ISL_452850\|2020- | .................... | .................... | ..................... | .................... | .................... | .................... |
| C | 8303 | hCoV-19/USA/VA-DCLS-0282/2020\|EPI_ISL_452851\|2020- | .................... | .................... | ..................... | .................... | .................... | .................... |
| C | 8304 | hCoV-19/USA/VA-DCLS-0283/2020\|EPI_ISL_452852\|2020- | .................... | .................... | ..................... | .................... | .................... | .................... |
| C | 8305 | hCoV-19/USA/VA-DCLS-0284/2020\|EPI_ISL_452853\|2020- | .................... | .................... | ..................... | .................... | .................... | .................... |
| C | 8306 | hCoV-19/USA/VA-DCLS-0285/2020\|EPI_ISL_452854\|2020- | .................... | .................... | ..................... | .................... | .................... | .................... |
| C | 8307 | hCoV-19/USA/VA-DCLS-0286/2020\|EPI_ISL_452855\|2020- | .................... | .................... | ..................... | .................... | .................... | .................... |
| C | 8308 | hCoV-19/USA/VA-DCLS-0287/2020\|EPI_ISL_452856\|2020- | .................... | .................... | ..................... | .................... | .................... | .................... |
| C | 8309 | hCoV-19/England/CAMB-1AED57/2020\|EPI_ISL_452857\|20 | .................... | .................... | ..................... | .................... | .................... | .................... |
| C | 8310 | hCoV-19/England/CAMB-1AED66/2020\|EPI_ISL_452858\|20 | .................... | .................... | ..................... | .................... | .................... | .................... |
| C | 8311 | hCoV-19/England/CAMB-1AED75/2020\|EPI_ISL_452859\|20 | .................... | .................... | ..................... | .................... | .................... | .................... |
| C | 8313 | hCoV-19/England/CAMB-1AEDB1/2020\|EPI_ISL_452863\|20 | .................... | .................... | ..................... | .................... | .................... | .................... |
| C | 8314 | hCoV-19/England/CAMB-1AEDC0/2020\|EPI_ISL_452864\|20 | .................... | .................... | ..................... | .................... | .................... | .................... |
| C | 8315 | hCoV-19/England/CAMB-1AEDDF/2020\|EPI_ISL_452865\|20 | .................... | .................... | ..................... | .................... | .................... | .................... |
| C | 8316 | hCoV-19/England/CAMB-1AEDEE/2020\|EPI_ISL_452866\|20 | .................... | .................... | ..................... | .................... | .................... | .................... |
| C | 8317 | hCoV-19/England/CAMB-1AEE54/2020\|EPI_ISL_452867\|20 | .................... | .................... | ..................... | .................... | .................... | .................... |
| C | 8318 | hCoV-19/England/CAMB-1AEEBE/2020\|EPI_ISL_452869\|20 | .................... | .................... | ..................... | .................... | .................... | .................... |
| C | 8319 | hCoV-19/England/CAMB-1AEF15/2020\|EPI_ISL_452871\|20 | .................... | .................... | ..................... | .................... | .................... | .................... |
| C | 8320 | hCoV-19/India/GJ-GBRC80/2020\|EPI_ISL_455015\|2020-0 | .................... | .................... | ..................... | .................... | .................... | .................... |
| C | 8321 | hCoV-19/India/GJ-GBRC119/2020\|EPI_ISL_455016\|2020- | .................... | .................... | ..................... | .................... | .................... | .................... |
| C | 8322 | hCoV-19/India/GJ-GBRC120/2020\|EPI_ISL_455017\|2020- | .................... | .................... | ..................... | .................... | .................... | .................... |
| C | 8323 | hCoV-19/India/GJ-GBRC121/2020\|EPI_ISL_455018\|2020- | .................... | .................... | ..................... | .................... | .................... | .................... |
| C | 8324 | hCoV-19/India/GJ-GBRC122/2020\|EPI_ISL_455019\|2020- | .................... | .................... | ..................... | .................... | .................... | .................... |
| C | 8325 | hCoV-19/India/GJ-GBRC123/2020\|EPI_ISL_455020\|2020- | .................... | .................... | ..................... | .................... | .................... | .................... |
| C | 8326 | hCoV-19/India/GJ-GBRC124/2020\|EPI_ISL_455021\|2020- | .................... | .................... | ..................... | .................... | .................... | .................... |
| C | 8327 | hCoV-19/India/GJ-GBRC125/2020\|EPI_ISL_455022\|2020- | .................... | .................... | ..................... | .................... | .................... | .................... |
| C | 8328 | hCoV-19/India/GJ-GBRC126/2020\|EPI_ISL_455023\|2020- | .................... | .................... | ..................... | .................... | .................... | .................... |
| C | 8329 | hCoV-19/India/GJ-GBRC127/2020\|EPI_ISL_455024\|2020- | .................... | .................... | ..................... | .................... | .................... | .................... |
| C | 8330 | hCoV-19/India/GJ-GBRC128/2020\|EPI_ISL_455025\|2020- | .................... | .................... | ..................... | .................... | .................... | .................... |
| C | 8331 | hCoV-19/India/GJ-GBRC129/2020\|EPI_ISL_455026\|2020- | .................... | .................... | ..................... | .................... | .................... | .................... |
| C | 8332 | hCoV-19/India/GJ-GBRC130/2020\|EPI_ISL_455027\|2020- | .................... | .................... | ..................... | .................... | .................... | .................... |
| C | 8333 | hCoV-19/Australia/NSW520/2020\|EPI_ISL_455060\|2020- | .................... | .................... | ..................... | .................... | .................... | .................... |
| C | 8334 | hCoV-19/Australia/NSW521/2020\|EPI_ISL_455061\|2020- | .................... | .................... | ..................... | .................... | .................... | .................... |
| C | 8335 | hCoV-19/Australia/NSW522/2020\|EPI_ISL_455062\|2020- | .................... | .................... | ..................... | .................... | .................... | .................... |
| C | 8336 | hCoV-19/Australia/NSW523/2020\|EPI_ISL_455063\|2020- | .................... | .................... | ..................... | .................... | .................... | .................... |
| C | 8337 | hCoV-19/Australia/NSW524/2020\|EPI_ISL_455064\|2020- | .................... | .................... | ..................... | .................... | .................... | .................... |
| C | 8338 | hCoV-19/Australia/NSW525/2020\|EPI_ISL_455065\|2020- | .................... | .................... | ..................... | .................... | .................... | .................... |
| C | 8339 | hCoV-19/Australia/NSW527/2020\|EPI_ISL_455066\|2020- | .................... | .................... | ..................... | .................... | .................... | .................... |
| C | 8340 | hCoV-19/Australia/NSW528/2020\|EPI_ISL_455067\|2020- | .................... | .................... | ..................... | .................... | .................... | .................... |
| C | 8341 | hCoV-19/Australia/NSW529/2020\|EPI_ISL_455068\|2020- | .................... | .................... | ..................... | .................... | .................... | .................... |
| C | 8342 | hCoV-19/Australia/NSW530/2020\|EPI_ISL_455069\|2020- | .................... | .................... | ..................... | .................... | .................... | .................... |
| C | 8343 | hCoV-19/Australia/NSW531/2020\|EPI_ISL_455070\|2020- | .................... | .................... | ..................... | .................... | .................... | .................... |
| C | 8344 | hCoV-19/Australia/NSW532/2020\|EPI_ISL_455071\|2020- | .................... | .................... | ..................... | .................... | .................... | .................... |
| C | 8345 | hCoV-19/Australia/NSW533/2020\|EPI_ISL_455072\|2020- | .................... | .................... | ..................... | .................... | .................... | .................... |
| C | 8346 | hCoV-19/Australia/NSW534/2020\|EPI_ISL_455073\|2020- | .................... | .................... | ..................... | .................... | .................... | .................... |
| C | 8347 | hCoV-19/Australia/NSW535/2020\|EPI_ISL_455074\|2020- | .................... | .................... | ..................... | .................... | .................... | .................... |
| C | 8348 | hCoV-19/Australia/NSW536/2020\|EPI_ISL_455075\|2020- | .................... | .................... | ..................... | .................... | .................... | .................... |
| C | 8349 | hCoV-19/Australia/NSW537/2020\|EPI_ISL_455076\|2020- | .................... | .................... | ..................... | .................... | .................... | .................... |
| C | 8350 | hCoV-19/Australia/NSW538/2020\|EPI_ISL_455077\|2020- | .................... | .................... | ..................... | .................... | .................... | .................... |
| C | 8351 | hCoV-19/Australia/NSW539/2020\|EPI_ISL_455078\|2020- | .................... | .................... | ..................... | .................... | .................... | .................... |
| C | 8352 | hCoV-19/Australia/NSW540/2020\|EPI_ISL_455079\|2020- | .................... | .................... | ..................... | .................... | .................... | .................... |
| C | 8353 | hCoV-19/Australia/NSW541/2020\|EPI_ISL_455080\|2020- | .................... | .................... | ..................... | .................... | .................... | .................... |

| | | | | | | | |
|---|---|---|---|---|---|---|---|
| C | 8354 hCoV-19/Australia/NSW542/2020\|EPI_ISL_455081\|2020- | ···················· | ···················· | ···················· | ···················· | ···················· | ···················· |
| C | 8355 hCoV-19/Australia/NSW543/2020\|EPI_ISL_455082\|2020- | ···················· | ···················· | ···················· | ···················· | ···················· | ···················· |
| C | 8356 hCoV-19/Australia/NSW544/2020\|EPI_ISL_455083\|2020- | ···················· | ···················· | ···················· | ···················· | ···················· | ···················· |
| C | 8357 hCoV-19/Australia/NSW545/2020\|EPI_ISL_455084\|2020- | ···················· | ···················· | ···················· | ···················· | ···················· | ···················· |
| C | 8358 hCoV-19/Australia/NSW546/2020\|EPI_ISL_455085\|2020- | ···················· | ···················· | ···················· | ···················· | ···················· | ···················· |
| C | 8359 hCoV-19/Australia/NSW547/2020\|EPI_ISL_455086\|2020- | ···················· | ···················· | ···················· | ···················· | ···················· | ···················· |
| C | 8360 hCoV-19/Australia/NSW548/2020\|EPI_ISL_455087\|2020- | ···················· | ···················· | ···················· | ···················· | ···················· | ···················· |
| C | 8361 hCoV-19/Australia/NSW549/2020\|EPI_ISL_455088\|2020- | ···················· | ···················· | ···················· | ···················· | ···················· | ···················· |
| C | 8362 hCoV-19/Australia/NSW550/2020\|EPI_ISL_455089\|2020- | ···················· | ···················· | ···················· | ···················· | ···················· | ···················· |
| C | 8363 hCoV-19/Australia/NSW551/2020\|EPI_ISL_455090\|2020- | ···················· | ···················· | ···················· | ···················· | ···················· | ···················· |
| C | 8364 hCoV-19/Australia/NSW552/2020\|EPI_ISL_455091\|2020- | ···················· | ·A·················· | ···················· | ···················· | ···················· | ···················· |
| C | 8365 hCoV-19/Australia/NSW553/2020\|EPI_ISL_455092\|2020- | ···················· | ···················· | ···················· | ···················· | ···················· | ···················· |
| C | 8366 hCoV-19/Australia/NSW554/2020\|EPI_ISL_455093\|2020- | ···················· | ···················· | ···················· | ···················· | ···················· | ···················· |
| C | 8367 hCoV-19/Australia/NSW555/2020\|EPI_ISL_455094\|2020- | ···················· | ·A·················· | ···················· | ···················· | ···················· | ···················· |
| C | 8368 hCoV-19/Australia/NSW556/2020\|EPI_ISL_455095\|2020- | ···················· | ···················· | ···················· | ···················· | ···················· | ···················· |
| C | 8369 hCoV-19/Australia/NSW557/2020\|EPI_ISL_455096\|2020- | ···················· | ···················· | ···················· | ···················· | ···················· | ···················· |
| C | 8370 hCoV-19/Australia/NSW558/2020\|EPI_ISL_455097\|2020- | ···················· | ···················· | ···················· | ···················· | ···················· | ···················· |
| C | 8371 hCoV-19/Australia/NSW559/2020\|EPI_ISL_455098\|2020- | ···················· | ···················· | ···················· | ···················· | ···················· | ···················· |
| C | 8372 hCoV-19/Australia/NSW560/2020\|EPI_ISL_455099\|2020- | ···················· | ···················· | ···················· | ···················· | ···················· | ···················· |
| C | 8373 hCoV-19/New Zealand/20VR1936/2020\|EPI_ISL_456315\|2 | ···················· | ···················· | ···················· | ···················· | ···················· | ···················· |
| C | 8374 hCoV-19/New Zealand/20VR1937/2020\|EPI_ISL_456316\|2 | ···················· | ···················· | ···················· | ···················· | ···················· | ···················· |
| C | 8375 hCoV-19/New Zealand/20VR1941/2020\|EPI_ISL_456317\|2 | ···················· | ···················· | ···················· | ···················· | ···················· | ···················· |
| C | 8376 hCoV-19/New Zealand/20VR1949/2020\|EPI_ISL_456319\|2 | ···················· | ···················· | ···················· | ···················· | ···················· | ···················· |
| C | 8377 hCoV-19/New Zealand/20VR1950/2020\|EPI_ISL_456320\|2 | ···················· | ···················· | ···················· | ···················· | ···················· | ···················· |
| C | 8378 hCoV-19/New Zealand/20VR1984/2020\|EPI_ISL_456321\|2 | ···················· | ···················· | ···················· | ···················· | ···················· | ···················· |
| C | 8379 hCoV-19/New Zealand/20VR1985/2020\|EPI_ISL_456322\|2 | ···················· | ···················· | ···················· | ···················· | ···················· | ···················· |
| C | 8380 hCoV-19/New Zealand/20VR1990/2020\|EPI_ISL_456324\|2 | ···················· | ···················· | ···················· | ···················· | ···················· | ···················· |
| C | 8381 hCoV-19/New Zealand/20VR1997/2020\|EPI_ISL_456326\|2 | ···················· | ···················· | ···················· | ···················· | ···················· | ···················· |
| C | 8382 hCoV-19/New Zealand/20VR2000/2020\|EPI_ISL_456327\|2 | ···················· | ···················· | ···················· | ···················· | ···················· | ···················· |
| C | 8383 hCoV-19/New Zealand/20VR2001/2020\|EPI_ISL_456328\|2 | ···················· | ···················· | ···················· | ···················· | ···················· | ···················· |
| C | 8384 hCoV-19/New Zealand/20VR2002/2020\|EPI_ISL_456329\|2 | ···················· | ···················· | ···················· | ···················· | ···················· | ···················· |
| C | 8385 hCoV-19/New Zealand/20VR2004/2020\|EPI_ISL_456330\|2 | ···················· | ···················· | ···················· | ···················· | ···················· | ···················· |
| C | 8386 hCoV-19/New Zealand/20VR2006/2020\|EPI_ISL_456331\|2 | ···················· | ···················· | ···················· | ···················· | ···················· | ···················· |
| C | 8387 hCoV-19/New Zealand/20VR2011/2020\|EPI_ISL_456332\|2 | ···················· | ···················· | ···················· | ···················· | ···················· | ···················· |
| C | 8388 hCoV-19/New Zealand/20VR2015/2020\|EPI_ISL_456334\|2 | ···················· | ···················· | ···················· | ···················· | ···················· | ···················· |
| C | 8389 hCoV-19/New Zealand/20VR2017/2020\|EPI_ISL_456335\|2 | ···················· | ···················· | ···················· | ···················· | ···················· | ···················· |
| C | 8390 hCoV-19/New Zealand/20VR2019/2020\|EPI_ISL_456336\|2 | ···················· | ···················· | ···················· | ···················· | ···················· | ···················· |
| C | 8391 hCoV-19/New Zealand/20VR2030/2020\|EPI_ISL_456338\|2 | ···················· | ···················· | ···················· | ···················· | ···················· | ···················· |
| C | 8392 hCoV-19/New Zealand/20VR2032/2020\|EPI_ISL_456339\|2 | ···················· | ···················· | ···················· | ···················· | ···················· | ···················· |
| C | 8393 hCoV-19/New Zealand/20VR2034/2020\|EPI_ISL_456340\|2 | ···················· | ······N············· | ···················· | ···················· | ···················· | ···················· |
| C | 8394 hCoV-19/New Zealand/20VR2037/2020\|EPI_ISL_456342\|2 | ···················· | ···················· | ···················· | ···················· | ···················· | ···················· |
| C | 8395 hCoV-19/New Zealand/20VR2045/2020\|EPI_ISL_456344\|2 | ···················· | ···················· | ···················· | ···················· | ···················· | ···················· |
| C | 8396 hCoV-19/New Zealand/20VR2049/2020\|EPI_ISL_456347\|2 | ···················· | ···················· | ···················· | ···················· | ···················· | ···················· |
| C | 8397 hCoV-19/New Zealand/20VR2051/2020\|EPI_ISL_456348\|2 | ···················· | ···················· | ···················· | ···················· | ···················· | ···················· |
| C | 8398 hCoV-19/New Zealand/20VR2058/2020\|EPI_ISL_456352\|2 | ···················· | ···················· | ···················· | ···················· | ···················· | ···················· |
| C | 8399 hCoV-19/New Zealand/20VR2059/2020\|EPI_ISL_456353\|2 | ···················· | ···················· | ···················· | ···················· | ···················· | ···················· |
| C | 8400 hCoV-19/New Zealand/20VR2063/2020\|EPI_ISL_456355\|2 | ···················· | ···················· | ···················· | ···················· | ···················· | ···················· |
| C | 8401 hCoV-19/New Zealand/20VR2065/2020\|EPI_ISL_456356\|2 | ···················· | ···················· | ···················· | ···················· | ···················· | ···················· |
| C | 8402 hCoV-19/New Zealand/20VR2067/2020\|EPI_ISL_456358\|2 | ···················· | ···················· | ···················· | ···················· | ···················· | ···················· |
| C | 8403 hCoV-19/New Zealand/20VR2556/2020\|EPI_ISL_456386\|2 | ···················· | ···················· | ···················· | ···················· | ···················· | ···················· |
| C | 8404 hCoV-19/New Zealand/20VR2917/2020\|EPI_ISL_456390\|2 | ···················· | ···················· | ···················· | ···················· | ···················· | ···················· |
| C | 8405 hCoV-19/New Zealand/20VR2932/2020\|EPI_ISL_456393\|2 | ···················· | ···················· | ···················· | ···················· | ···················· | ···················· |
| C | 8406 hCoV-19/New Zealand/20VR2941/2020\|EPI_ISL_456394\|2 | ···················· | ···················· | ···················· | ···················· | ···················· | ···················· |
| C | 8407 hCoV-19/New Zealand/20VR3003/2020\|EPI_ISL_456395\|2 | ···················· | ···················· | ···················· | ···················· | ···················· | ···················· |
| C | 8408 hCoV-19/New Zealand/20VR3004/2020\|EPI_ISL_456396\|2 | ···················· | ···················· | ···················· | ···················· | ···················· | ···················· |
| C | 8409 hCoV-19/New Zealand/20VR3005/2020\|EPI_ISL_456397\|2 | ···················· | ···················· | ···················· | ···················· | ···················· | ···················· |
| C | 8410 hCoV-19/England/SHEF-CB7C0/2020\|EPI_ISL_457647\|202 | ···················· | ···················· | ···················· | ···················· | ···················· | ···················· |
| C | 8411 hCoV-19/England/SHEF-CB809/2020\|EPI_ISL_457648\|202 | ···················· | ···················· | ···················· | ···················· | ···················· | ···················· |
| C | 8412 hCoV-19/England/SHEF-CC3DA/2020\|EPI_ISL_457652\|202 | ···················· | ···················· | ···················· | ···················· | ···················· | ···················· |
| C | 8413 hCoV-19/England/SHEF-CC6E0/2020\|EPI_ISL_457654\|202 | ···················· | ···················· | ···················· | ···················· | ···················· | ···················· |
| C | 8414 hCoV-19/England/SHEF-CC817/2020\|EPI_ISL_457655\|202 | ···················· | ···················· | ···················· | ···················· | ···················· | ···················· |
| C | 8415 hCoV-19/England/SHEF-CC9E7/2020\|EPI_ISL_457657\|202 | ···················· | ···················· | ···················· | ···················· | ···················· | ···················· |
| C | 8416 hCoV-19/England/SHEF-CCFB8/2020\|EPI_ISL_457658\|202 | ···················· | ···················· | ···················· | ···················· | ···················· | ···················· |
| C | 8417 hCoV-19/England/SHEF-CDA2F/2020\|EPI_ISL_457659\|202 | ···················· | ···················· | ···················· | ···················· | ···················· | ···················· |
| C | 8418 hCoV-19/England/SHEF-CDBE0/2020\|EPI_ISL_457660\|202 | ···················· | ···················· | ···················· | ···················· | ···················· | ···················· |
| C | 8419 hCoV-19/England/SHEF-CDC29/2020\|EPI_ISL_457661\|202 | ···················· | ···················· | ···················· | ···················· | ···················· | ···················· |
| C | 8420 hCoV-19/England/SHEF-CEE8C/2020\|EPI_ISL_457667\|202 | ···················· | ···················· | ···················· | ···················· | ···················· | ···················· |
| C | 8421 hCoV-19/England/SHEF-CFFF1/2020\|EPI_ISL_457670\|202 | ···················· | ···················· | ···················· | ···················· | ···················· | ···················· |
| C | 8422 hCoV-19/England/SHEF-D0388/2020\|EPI_ISL_457672\|202 | ···················· | ···················· | ···················· | ···················· | ···················· | ···················· |
| C | 8423 hCoV-19/England/SHEF-D07AA/2020\|EPI_ISL_457675\|202 | ···················· | ···················· | ···················· | ···················· | ···················· | ···················· |
| C | 8424 hCoV-19/England/SHEF-D087A/2020\|EPI_ISL_457676\|202 | ···················· | ···················· | ···················· | ···················· | ···················· | ···················· |
| C | 8425 hCoV-19/England/SHEF-D0F57/2020\|EPI_ISL_457679\|202 | ···················· | ···················· | ···················· | ···················· | ···················· | ···················· |
| C | 8426 hCoV-19/England/SHEF-D0F75/2020\|EPI_ISL_457680\|202 | ···················· | ···················· | ···················· | ···················· | ···················· | ···················· |
| C | 8427 hCoV-19/England/SHEF-D1C22/2020\|EPI_ISL_457683\|202 | ···················· | ···················· | ···················· | ···················· | ···················· | ···················· |
| C | 8428 hCoV-19/England/SHEF-D2EA3/2020\|EPI_ISL_457685\|202 | ···················· | ···················· | ···················· | ···················· | ···················· | ···················· |
| C | 8429 hCoV-19/Guangzhou/GZMU0074/2020\|EPI_ISL_457687\|202 | ···················· | ···················· | ···················· | ···················· | ···················· | ···················· |
| C | 8430 hCoV-19/Guangzhou/GZMU0075/2020\|EPI_ISL_457688\|202 | ···················· | ···················· | ···················· | ···················· | ···················· | ···················· |
| C | 8431 hCoV-19/Guangzhou/GZMU0077/2020\|EPI_ISL_457689\|202 | ···················· | ···················· | ···················· | ···················· | ···················· | ···················· |

| | | | | | | | | |
|---|---|---|---|---|---|---|---|---|
| C | 8432 | hCoV-19/England/BRIS-126CE1/2020\|EPI_ISL_458977\|20 | .................... | .................... | .................... | .................... | .................... | .................... |
| C | 8433 | hCoV-19/England/BRIS-126BE4/2020\|EPI_ISL_458978\|20 | .................... | .................... | .................... | .................... | .................... | .................... |
| C | 8434 | hCoV-19/England/BRIS-12663E/2020\|EPI_ISL_458979\|20 | .................... | .................... | .................... | .................... | .................... | .................... |
| C | 8435 | hCoV-19/England/BRIS-126698/2020\|EPI_ISL_458981\|20 | .................... | .................... | .................... | .................... | .................... | .................... |
| C | 8436 | hCoV-19/England/BRIS-126BA8/2020\|EPI_ISL_458982\|20 | .................... | .................... | .................... | .................... | .................... | .................... |
| C | 8437 | hCoV-19/England/BRIS-126294/2020\|EPI_ISL_458983\|20 | .................... | .................... | .................... | .................... | .................... | .................... |
| C | 8438 | hCoV-19/England/BRIS-126AF6/2020\|EPI_ISL_458984\|20 | .................... | .................... | .................... | .................... | .................... | .................... |
| C | 8439 | hCoV-19/England/BRIS-126391/2020\|EPI_ISL_458985\|20 | .................... | .................... | .................... | .................... | .................... | .................... |
| C | 8440 | hCoV-19/England/BRIS-12607C/2020\|EPI_ISL_458986\|20 | .................... | .................... | .................... | .................... | .................... | .................... |
| C | 8441 | hCoV-19/England/BRIS-126328/2020\|EPI_ISL_458987\|20 | .................... | .................... | .................... | .................... | .................... | .................... |
| C | 8442 | hCoV-19/England/BRIS-126CD2/2020\|EPI_ISL_458989\|20 | .................... | .................... | .................... | .................... | .................... | .................... |
| C | 8443 | hCoV-19/England/BRIS-126434/2020\|EPI_ISL_458990\|20 | .................... | .................... | .................... | .................... | .................... | .................... |
| C | 8444 | hCoV-19/England/BRIS-126BF3/2020\|EPI_ISL_458991\|20 | .................... | .................... | .................... | .................... | .................... | .................... |
| C | 8445 | hCoV-19/England/BRIS-1268ED/2020\|EPI_ISL_458992\|20 | .................... | .................... | .................... | .................... | .................... | .................... |
| C | 8446 | hCoV-19/England/BRIS-1263FB/2020\|EPI_ISL_458994\|20 | .................... | .................... | .................... | .................... | .................... | .................... |
| C | 8447 | hCoV-19/England/BRIS-126838/2020\|EPI_ISL_458995\|20 | .................... | .................... | .................... | .................... | .................... | .................... |
| C | 8448 | hCoV-19/England/BRIS-126C3C/2020\|EPI_ISL_458998\|20 | .................... | .................... | .................... | .................... | .................... | .................... |
| C | 8449 | hCoV-19/Uganda/UG019/2020\|EPI_ISL_451201\|2020-04-3 | .................... | .................... | .................... | .................... | .................... | .................... |
| C | 8450 | hCoV-19/Uganda/UG020/2020\|EPI_ISL_451202\|2020-05-0 | .................... | .................... | .................... | .................... | .................... | .................... |
| C | 8451 | hCoV-19/USA/LA-EVTL042/2020\|EPI_ISL_451203\|2020-05 | .................... | .................... | .................... | .................... | .................... | .................... |
| C | 8452 | hCoV-19/USA/LA-EVTL043/2020\|EPI_ISL_451204\|2020-04 | .................... | .................... | .................... | .................... | .................... | .................... |
| C | 8453 | hCoV-19/USA/LA-EVTL044/2020\|EPI_ISL_451205\|2020-05 | .................... | .................... | .................... | .................... | .................... | .................... |
| C | 8454 | hCoV-19/USA/LA-EVTL045/2020\|EPI_ISL_451206\|2020-05 | .................... | .................... | .................... | .................... | .................... | .................... |
| C | 8455 | hCoV-19/USA/LA-EVTL046/2020\|EPI_ISL_451207\|2020-05 | .................... | .................... | .................... | .................... | .................... | .................... |
| C | 8456 | hCoV-19/USA/LA-EVTL047/2020\|EPI_ISL_451208\|2020-05 | .................... | .................... | .................... | .................... | .................... | .................... |
| C | 8457 | hCoV-19/USA/LA-EVTL048/2020\|EPI_ISL_451209\|2020-05 | .................... | .................... | .................... | .................... | .................... | .................... |
| C | 8458 | hCoV-19/USA/LA-EVTL049/2020\|EPI_ISL_451210\|2020-05 | .................... | .................... | .................... | .................... | .................... | .................... |
| C | 8459 | hCoV-19/USA/LA-EVTL050/2020\|EPI_ISL_451211\|2020-05 | .................... | .A.................. | .................... | .................... | .................... | .................... |
| C | 8460 | hCoV-19/USA/LA-EVTL051/2020\|EPI_ISL_451212\|2020-04 | .................... | .................... | .................... | .................... | .................... | .................... |
| C | 8461 | hCoV-19/USA/LA-EVTL052/2020\|EPI_ISL_451213\|2020-05 | .................... | .................... | .................... | .................... | .................... | .................... |
| C | 8462 | hCoV-19/USA/LA-EVTL053/2020\|EPI_ISL_451214\|2020-04 | .................... | .................... | .................... | .................... | .................... | .................... |
| C | 8463 | hCoV-19/USA/LA-EVTL054/2020\|EPI_ISL_451215\|2020-04 | .................... | .................... | .................... | .................... | .................... | .................... |
| C | 8464 | hCoV-19/USA/LA-EVTL055/2020\|EPI_ISL_451216\|2020-04 | .................... | .................... | .................... | .................... | .................... | .................... |
| C | 8465 | hCoV-19/USA/LA-EVTL056/2020\|EPI_ISL_451217\|2020-05 | .................... | .................... | .................... | .................... | .................... | .................... |
| C | 8466 | hCoV-19/USA/LA-EVTL057/2020\|EPI_ISL_451218\|2020-04 | .................... | .................... | .................... | .................... | .................... | .................... |
| C | 8467 | hCoV-19/USA/LA-EVTL058/2020\|EPI_ISL_451219\|2020-04 | .................... | .................... | .................... | .................... | .................... | .................... |
| C | 8468 | hCoV-19/USA/LA-EVTL059/2020\|EPI_ISL_451220\|2020-04 | .................... | .................... | .................... | .................... | .................... | .................... |
| C | 8469 | hCoV-19/USA/LA-EVTL060/2020\|EPI_ISL_451221\|2020-04 | .................... | .................... | .................... | .................... | .................... | .................... |
| C | 8470 | hCoV-19/USA/LA-EVTL072/2020\|EPI_ISL_451233\|2020-05 | .................... | .................... | .................... | .................... | .................... | .................... |
| C | 8471 | hCoV-19/USA/LA-EVTL073/2020\|EPI_ISL_451234\|2020-05 | .................... | .................... | .................... | .................... | .................... | .................... |
| C | 8472 | hCoV-19/USA/LA-EVTL074/2020\|EPI_ISL_451235\|2020-05 | .................... | .................... | .................... | .................... | .................... | .................... |
| C | 8473 | hCoV-19/USA/LA-EVTL075/2020\|EPI_ISL_451236\|2020-05 | .................... | .................... | .................... | .................... | .................... | .................... |
| C | 8474 | hCoV-19/USA/LA-EVTL076/2020\|EPI_ISL_451237\|2020-05 | .................... | .................... | .................... | .................... | .................... | .................... |
| C | 8475 | hCoV-19/USA/LA-EVTL077/2020\|EPI_ISL_451238\|2020-05 | .................... | .................... | .................... | .................... | .................... | .................... |
| C | 8476 | hCoV-19/USA/LA-EVTL083/2020\|EPI_ISL_451244\|2020-05 | .................... | .................... | .................... | .................... | .................... | .................... |
| C | 8477 | hCoV-19/USA/LA-EVTL084/2020\|EPI_ISL_451245\|2020-05 | .................... | .................... | .................... | .................... | .................... | .................... |
| C | 8478 | hCoV-19/USA/LA-EVTL085/2020\|EPI_ISL_451246\|2020-05 | .................... | .................... | .................... | .................... | .................... | .................... |
| C | 8479 | hCoV-19/USA/LA-EVTL086/2020\|EPI_ISL_451247\|2020-05 | .................... | .................... | .................... | .................... | .................... | .................... |
| C | 8480 | hCoV-19/USA/LA-EVTL087/2020\|EPI_ISL_451248\|2020-05 | .................... | .................... | .................... | .................... | .................... | .................... |
| C | 8481 | hCoV-19/USA/LA-EVTL088/2020\|EPI_ISL_451249\|2020-05 | .................... | .................... | .................... | .................... | .................... | .................... |
| C | 8482 | hCoV-19/USA/LA-EVTL089/2020\|EPI_ISL_451250\|2020-05 | .................... | .................... | .................... | .................... | .................... | .................... |
| C | 8483 | hCoV-19/USA/LA-EVTL090/2020\|EPI_ISL_451251\|2020-05 | .................... | .................... | .................... | .................... | .................... | .................... |
| C | 8484 | hCoV-19/USA/LA-EVTL091/2020\|EPI_ISL_451252\|2020-05 | .................... | .................... | .................... | .................... | .................... | .................... |
| C | 8485 | hCoV-19/USA/LA-EVTL092/2020\|EPI_ISL_451253\|2020-05 | .................... | .................... | .................... | .................... | .................... | .................... |
| C | 8486 | hCoV-19/USA/LA-EVTL093/2020\|EPI_ISL_451254\|2020-05 | .................... | .................... | .................... | .................... | .................... | .................... |
| C | 8487 | hCoV-19/USA/LA-EVTL094/2020\|EPI_ISL_451255\|2020-05 | .................... | .................... | .................... | .................... | .................... | .................... |
| C | 8488 | hCoV-19/USA/LA-EVTL095/2020\|EPI_ISL_451256\|2020-05 | .................... | .................... | .................... | .................... | .................... | .................... |
| C | 8489 | hCoV-19/USA/LA-EVTL096/2020\|EPI_ISL_451257\|2020-05 | .................... | .................... | .................... | .................... | .................... | .................... |
| C | 8490 | hCoV-19/USA/LA-EVTL097/2020\|EPI_ISL_451258\|2020-05 | .................... | .................... | .................... | .................... | .................... | .................... |
| C | 8491 | hCoV-19/USA/LA-EVTL098/2020\|EPI_ISL_451259\|2020-05 | .................... | .................... | .................... | .................... | .................... | .................... |
| C | 8492 | hCoV-19/USA/LA-EVTL099/2020\|EPI_ISL_451260\|2020-05 | .................... | .................... | .................... | .................... | .................... | .................... |
| C | 8493 | hCoV-19/USA/LA-EVTL100/2020\|EPI_ISL_451261\|2020-05 | .................... | .................... | .................... | .................... | .................... | .................... |
| C | 8494 | hCoV-19/USA/LA-EVTL101/2020\|EPI_ISL_451262\|2020-05 | .................... | .................... | .................... | .................... | .................... | .................... |
| C | 8495 | hCoV-19/USA/LA-EVTL102/2020\|EPI_ISL_451263\|2020-05 | .................... | .................... | .................... | .................... | .................... | .................... |
| C | 8496 | hCoV-19/USA/LA-EVTL103/2020\|EPI_ISL_451264\|2020-05 | .................... | .................... | .................... | .................... | .................... | .................... |
| C | 8497 | hCoV-19/USA/LA-EVTL104/2020\|EPI_ISL_451265\|2020-05 | .................... | .................... | .................... | .................... | .................... | .................... |
| C | 8498 | hCoV-19/USA/LA-EVTL105/2020\|EPI_ISL_451266\|2020-05 | .................... | .................... | .................... | .................... | .................... | .................... |
| C | 8499 | hCoV-19/USA/LA-EVTL106/2020\|EPI_ISL_451267\|2020-05 | .................... | .................... | .................... | .................... | .................... | .................... |
| C | 8500 | hCoV-19/USA/LA-EVTL107/2020\|EPI_ISL_451268\|2020-05 | .................... | .................... | .................... | .................... | .................... | .................... |
| C | 8501 | hCoV-19/USA/LA-EVTL108/2020\|EPI_ISL_451269\|2020-05 | .................... | .................... | .................... | .................... | .................... | .................... |
| C | 8502 | hCoV-19/USA/LA-EVTL109/2020\|EPI_ISL_451270\|2020-05 | .................... | .................... | .................... | .................... | .................... | .................... |
| C | 8503 | hCoV-19/USA/LA-EVTL110/2020\|EPI_ISL_451271\|2020-05 | .................... | .................... | .................... | .................... | .................... | .................... |
| C | 8504 | hCoV-19/USA/LA-EVTL111/2020\|EPI_ISL_451272\|2020-05 | .................... | .................... | .................... | .................... | .................... | .................... |
| C | 8505 | hCoV-19/USA/LA-EVTL112/2020\|EPI_ISL_451273\|2020-05 | .................... | .................... | .................... | .................... | .................... | .................... |
| C | 8506 | hCoV-19/USA/LA-EVTL113/2020\|EPI_ISL_451274\|2020-05 | .................... | .................... | .................... | .................... | .................... | .................... |
| C | 8507 | hCoV-19/USA/LA-EVTL114/2020\|EPI_ISL_451275\|2020-05 | .................... | .................... | .................... | .................... | .................... | .................... |
| C | 8508 | hCoV-19/USA/LA-EVTL115/2020\|EPI_ISL_451276\|2020-05 | .................... | .................... | .................... | .................... | .................... | .................... |
| C | 8509 | hCoV-19/USA/LA-EVTL116/2020\|EPI_ISL_451277\|2020-05 | .................... | .................... | .................... | .................... | .................... | .................... |

| | | | | | | | | |
|---|---|---|---|---|---|---|---|---|
| C | 8510 hCoV-19/USA/LA-EVTL117/2020\|EPI_ISL_451278\|2020-05 | ···················· | ···················· | ···················· | ···················· | ···················· | ···················· | |
| C | 8511 hCoV-19/USA/LA-EVTL118/2020\|EPI_ISL_451279\|2020-05 | ···················· | ···················· | ···················· | ···················· | ···················· | ···················· | |
| C | 8512 hCoV-19/USA/LA-EVTL119/2020\|EPI_ISL_451280\|2020-05 | ···················· | ···················· | ···················· | ···················· | ···················· | ···················· | |
| C | 8513 hCoV-19/USA/LA-EVTL120/2020\|EPI_ISL_451281\|2020-05 | ···················· | ···················· | ···················· | ···················· | ···················· | ···················· | |
| C | 8514 hCoV-19/USA/LA-EVTL121/2020\|EPI_ISL_451282\|2020-05 | ···················· | ···················· | ···················· | ···················· | ···················· | ···················· | |
| C | 8515 hCoV-19/USA/LA-EVTL122/2020\|EPI_ISL_451283\|2020-05 | ···················· | ···················· | ···················· | ···················· | ···················· | ···················· | |
| C | 8516 hCoV-19/USA/LA-EVTL123/2020\|EPI_ISL_451284\|2020-05 | ···················· | ···················· | ···················· | ···················· | ···················· | ···················· | |
| C | 8517 hCoV-19/USA/LA-EVTL124/2020\|EPI_ISL_451285\|2020-05 | ···················· | ···················· | ···················· | ···················· | ···················· | ···················· | |
| C | 8518 hCoV-19/USA/LA-EVTL125/2020\|EPI_ISL_451286\|2020-05 | ···················· | ···················· | ···················· | ···················· | ···················· | ···················· | |
| C | 8519 hCoV-19/USA/LA-EVTL126/2020\|EPI_ISL_451287\|2020-05 | ···················· | ···················· | ···················· | ···················· | ···················· | ···················· | |
| C | 8520 hCoV-19/USA/LA-EVTL127/2020\|EPI_ISL_451288\|2020-05 | ···················· | ···················· | ···················· | ···················· | ···················· | ···················· | |
| C | 8521 hCoV-19/USA/LA-EVTL128/2020\|EPI_ISL_451289\|2020-05 | ···················· | ···················· | ···················· | ···················· | ···················· | ···················· | |
| C | 8522 hCoV-19/USA/LA-EVTL129/2020\|EPI_ISL_451290\|2020-05 | ···················· | ···················· | ···················· | ···················· | ···················· | ···················· | |
| C | 8523 hCoV-19/USA/LA-EVTL130/2020\|EPI_ISL_451291\|2020-05 | ···················· | ···················· | ···················· | ···················· | ···················· | ···················· | |
| C | 8524 hCoV-19/USA/LA-EVTL131/2020\|EPI_ISL_451292\|2020-05 | ···················· | ···················· | ···················· | ···················· | ···················· | ···················· | |
| C | 8525 hCoV-19/USA/LA-EVTL132/2020\|EPI_ISL_451293\|2020-05 | ···················· | ···················· | ···················· | ···················· | ···················· | ···················· | |
| C | 8526 hCoV-19/USA/LA-EVTL133/2020\|EPI_ISL_451294\|2020-05 | ···················· | ···················· | ···················· | ···················· | ···················· | ···················· | |
| C | 8527 hCoV-19/USA/LA-EVTL134/2020\|EPI_ISL_451295\|2020-05 | ···················· | ···················· | ···················· | ···················· | ···················· | ···················· | |
| C | 8528 hCoV-19/USA/LA-EVTL135/2020\|EPI_ISL_451296\|2020-05 | ···················· | ···················· | ···················· | ···················· | ···················· | ···················· | |
| C | 8529 hCoV-19/USA/LA-EVTL136/2020\|EPI_ISL_451297\|2020-05 | ···················· | ···················· | ···················· | ···················· | ···················· | ···················· | |
| C | 8530 hCoV-19/Italy/LAZ-INMI1-B1/2020\|EPI_ISL_451298\|202 | ···················· | ···················· | ···················· | ···················· | ···················· | ···················· | |
| C | 8531 hCoV-19/Italy/LAZ-INMI1-B2/2020\|EPI_ISL_451299\|202 | ···················· | ···················· | ···················· | ···················· | ···················· | ···················· | |
| C | 8535 hCoV-19/Spain/NC-IBV-001385/2020\|EPI_ISL_452516\|20 | ···················· | ···················· | ···················· | ···················· | ···················· | ···················· | |
| C | 8539 hCoV-19/Spain/NC-IBV-001381/2020\|EPI_ISL_452520\|20 | ···················· | ···················· | ···················· | ···················· | ···················· | ···················· | |
| C | 8541 hCoV-19/Spain/NC-IBV-001378/2020\|EPI_ISL_452523\|20 | ···················· | ···················· | ···················· | ···················· | ···················· | ···················· | |
| C | 8546 hCoV-19/Spain/NC-IBV-001370/2020\|EPI_ISL_452531\|20 | ···················· | ···················· | ···················· | ···················· | ···················· | ···················· | |
| C | 8548 hCoV-19/Spain/MD-IBV-001367/2020\|EPI_ISL_452534\|20 | ···················· | ···················· | ···················· | ···················· | ···················· | ···················· | |
| C | 8549 hCoV-19/Spain/MD-IBV-001366/2020\|EPI_ISL_452535\|20 | ···················· | ···················· | ···················· | ···················· | ···················· | ···················· | |
| C | 8551 hCoV-19/Spain/NC-IBV-001362/2020\|EPI_ISL_452539\|20 | ···················· | ···················· | ···················· | ···················· | ·NNN················ | ···················· | 1 |
| C | 8552 hCoV-19/Spain/NC-IBV-001361/2020\|EPI_ISL_452540\|20 | ···················· | ···················· | ···················· | ···················· | ···················· | ···················· | |
| C | 8562 hCoV-19/Spain/AN-IBV-001176/2020\|EPI_ISL_452567\|20 | ···················· | ···················· | ···················· | ···················· | ···················· | ···················· | |
| C | 8565 hCoV-19/Spain/AN-IBV-001155/2020\|EPI_ISL_452577\|20 | ···················· | ···················· | ···················· | ···················· | ···················· | ···················· | |
| C | 8573 hCoV-19/Spain/AN-IBV-001104/2020\|EPI_ISL_452586\|20 | ···················· | ···················· | ···················· | ···················· | ···················· | ···················· | |
| C | 8577 hCoV-19/Spain/AN-IBV-001093/2020\|EPI_ISL_452593\|20 | ···················· | ···················· | ···················· | ···················· | ···················· | ···················· | |
| C | 8581 hCoV-19/Spain/AN-IBV-001089/2020\|EPI_ISL_452597\|20 | ···················· | ···················· | ···················· | ···················· | ···················· | ···················· | |
| C | 8582 hCoV-19/Spain/AN-IBV-001088/2020\|EPI_ISL_452598\|20 | ···················· | ···················· | ···················· | ···················· | ···················· | ···················· | |
| C | 8583 hCoV-19/Portugal/PT0124/2020\|EPI_ISL_453840\|2020-0 | ···················· | ···················· | ···················· | ···················· | ···················· | ···················· | |
| C | 8584 hCoV-19/Portugal/PT0128/2020\|EPI_ISL_453844\|2020-0 | ···················· | ···················· | ···················· | ···················· | ···················· | ···················· | |
| C | 8585 hCoV-19/Portugal/PT0129/2020\|EPI_ISL_453845\|2020-0 | ···················· | ···················· | ···················· | ···················· | ···················· | ···················· | |
| C | 8586 hCoV-19/Portugal/PT0130/2020\|EPI_ISL_453846\|2020-0 | ···················· | ···················· | ···················· | ···················· | ···················· | ···················· | |
| C | 8587 hCoV-19/Portugal/PT0131/2020\|EPI_ISL_453847\|2020-0 | ···················· | ···················· | ···················· | ···················· | ···················· | ···················· | |
| C | 8588 hCoV-19/Portugal/PT0132/2020\|EPI_ISL_453848\|2020-0 | ···················· | ···················· | ···················· | ···················· | ···················· | ···················· | |
| C | 8589 hCoV-19/Portugal/PT0134/2020\|EPI_ISL_453850\|2020-0 | ···················· | ···················· | ···················· | ···················· | ···················· | ···················· | |
| C | 8590 hCoV-19/Portugal/PT0135/2020\|EPI_ISL_453851\|2020-0 | ···················· | ···················· | ···················· | ···················· | ···················· | ···················· | |
| C | 8591 hCoV-19/Portugal/PT0136/2020\|EPI_ISL_453852\|2020-0 | ···················· | ···················· | ···················· | ···················· | ···················· | ···················· | |
| C | 8592 hCoV-19/Portugal/PT0137/2020\|EPI_ISL_453853\|2020-0 | ···················· | ···················· | ···················· | ···················· | ···················· | ···················· | |
| C | 8593 hCoV-19/Portugal/PT0138/2020\|EPI_ISL_453854\|2020-0 | ···················· | ···················· | ···················· | ···················· | ···················· | ···················· | |
| C | 8594 hCoV-19/Portugal/PT0139/2020\|EPI_ISL_453855\|2020-0 | ···················· | ···················· | ···················· | ···················· | ···················· | ···················· | |
| C | 8595 hCoV-19/Portugal/PT0140/2020\|EPI_ISL_453856\|2020-0 | ···················· | ···················· | ···················· | ···················· | ···················· | ···················· | |
| C | 8596 hCoV-19/Portugal/PT0143/2020\|EPI_ISL_453859\|2020-0 | ···················· | ···················· | ···················· | ···················· | ···················· | ···················· | |
| C | 8597 hCoV-19/Portugal/PT0144/2020\|EPI_ISL_453860\|2020-0 | ···················· | ···················· | ···················· | ···················· | ···················· | ···················· | |
| C | 8598 hCoV-19/Portugal/PT0149/2020\|EPI_ISL_453865\|2020-0 | ···················· | ···················· | ···················· | ···················· | ···················· | ···················· | |
| C | 8599 hCoV-19/Portugal/PT0150/2020\|EPI_ISL_453866\|2020-0 | ···················· | ···················· | ···················· | ···················· | ···················· | ···················· | |
| C | 8600 hCoV-19/Portugal/PT0152/2020\|EPI_ISL_453868\|2020-0 | ···················· | ···················· | ···················· | ···················· | ···················· | ···················· | |
| C | 8601 hCoV-19/Portugal/PT0153/2020\|EPI_ISL_453869\|2020-0 | ···················· | ···················· | ···················· | ···················· | ···················· | ···················· | |
| C | 8602 hCoV-19/Portugal/PT0154/2020\|EPI_ISL_453870\|2020-0 | ···················· | ···················· | ···················· | ···················· | ···················· | ···················· | |
| C | 8603 hCoV-19/Portugal/PT0156/2020\|EPI_ISL_453872\|2020-0 | ···················· | ···················· | ···················· | ···················· | ···················· | ···················· | |
| C | 8604 hCoV-19/Portugal/PT0157/2020\|EPI_ISL_453873\|2020-0 | ···················· | ···················· | ···················· | ···················· | ···················· | ···················· | |
| C | 8605 hCoV-19/Portugal/PT0159/2020\|EPI_ISL_453875\|2020-0 | ···················· | ···················· | ···················· | ···················· | ···················· | ···················· | |
| C | 8606 hCoV-19/Portugal/PT0160/2020\|EPI_ISL_453876\|2020-0 | ···················· | ···················· | ···················· | ···················· | ···················· | ···················· | |
| C | 8607 hCoV-19/Portugal/PT0161/2020\|EPI_ISL_453877\|2020-0 | ···················· | ···················· | ···················· | ···················· | ···················· | ···················· | |
| C | 8608 hCoV-19/Portugal/PT0163/2020\|EPI_ISL_453879\|2020-0 | ···················· | ···················· | ···················· | ···················· | ···················· | ···················· | |
| C | 8609 hCoV-19/Portugal/PT0165/2020\|EPI_ISL_453881\|2020-0 | ···················· | ···················· | ···················· | ···················· | ···················· | ···················· | |
| C | 8610 hCoV-19/Portugal/PT0179/2020\|EPI_ISL_453895\|2020-0 | ···················· | ···················· | ···················· | ···················· | ···················· | ···················· | |
| C | 8611 hCoV-19/Portugal/PT0180/2020\|EPI_ISL_453896\|2020-0 | ···················· | ···················· | ···················· | ···················· | ···················· | ···················· | |
| C | 8612 hCoV-19/Portugal/PT0182/2020\|EPI_ISL_453898\|2020-0 | ···················· | ···················· | ···················· | ···················· | ···················· | ···················· | |
| C | 8613 hCoV-19/Australia/NSW488/2020\|EPI_ISL_455028\|2020- | ···················· | ···················· | ···················· | ···················· | ···················· | ···················· | |
| C | 8614 hCoV-19/Australia/NSW489/2020\|EPI_ISL_455029\|2020- | ···················· | ···················· | ···················· | ···················· | ···················· | ···················· | |
| C | 8615 hCoV-19/Australia/NSW490/2020\|EPI_ISL_455030\|2020- | ···················· | ···················· | ···················· | ···················· | ···················· | ···················· | |
| C | 8616 hCoV-19/Australia/NSW491/2020\|EPI_ISL_455031\|2020- | ···················· | ···················· | ···········T········ | ···················· | ···················· | ···················· | |
| C | 8617 hCoV-19/Australia/NSW492/2020\|EPI_ISL_455032\|2020- | ···················· | ···················· | ···················· | ···················· | ···················· | ···················· | |
| C | 8618 hCoV-19/Australia/NSW493/2020\|EPI_ISL_455033\|2020- | ···················· | ···················· | ···················· | ···················· | ···················· | ···················· | |
| C | 8619 hCoV-19/Australia/NSW494/2020\|EPI_ISL_455034\|2020- | ···················· | ···················· | ···················· | ···················· | ···················· | ···················· | |
| C | 8620 hCoV-19/Australia/NSW495/2020\|EPI_ISL_455035\|2020- | ···················· | ···················· | ···················· | ···················· | ···················· | ···················· | |
| C | 8621 hCoV-19/Australia/NSW496/2020\|EPI_ISL_455036\|2020- | ···················· | ···················· | ···················· | ···················· | ···················· | ···················· | |
| C | 8622 hCoV-19/Australia/NSW497/2020\|EPI_ISL_455037\|2020- | ···················· | ···················· | ···················· | ···················· | ···················· | ···················· | |
| C | 8623 hCoV-19/Australia/NSW498/2020\|EPI_ISL_455038\|2020- | ···················· | ···················· | ···················· | ···················· | ···················· | ···················· | |
| C | 8624 hCoV-19/Australia/NSW499/2020\|EPI_ISL_455039\|2020- | ···················· | ···················· | ···················· | ···················· | ···················· | ···················· | |

| | | | | | | | | | |
|---|---|---|---|---|---|---|---|---|---|
| C | 8625 | hCoV-19/Australia/NSW500/2020\|EPI_ISL_455040\|2020- | ···················· | ···················· | ···················· | ···················· | ···················· | ···················· | |
| C | 8626 | hCoV-19/Australia/NSW501/2020\|EPI_ISL_455041\|2020- | ···················· | ···················· | ···················· | ···················· | ···················· | ···················· | |
| C | 8627 | hCoV-19/Australia/NSW502/2020\|EPI_ISL_455042\|2020- | ···················· | ···················· | ···················· | ···················· | ···················· | ···················· | |
| C | 8628 | hCoV-19/Australia/NSW503/2020\|EPI_ISL_455043\|2020- | ···················· | ·············A······ | ···················· | ···················· | ···················· | ···················· | |
| C | 8629 | hCoV-19/Australia/NSW504/2020\|EPI_ISL_455044\|2020- | ···················· | ···················· | ···················· | ···················· | ···················· | ···················· | |
| C | 8630 | hCoV-19/Australia/NSW505/2020\|EPI_ISL_455045\|2020- | ···················· | ···················· | ···················· | ···················· | ···················· | ···················· | |
| C | 8631 | hCoV-19/Australia/NSW506/2020\|EPI_ISL_455046\|2020- | ···················· | ···················· | ···················· | ···················· | ···················· | ···················· | |
| C | 8632 | hCoV-19/Australia/NSW507/2020\|EPI_ISL_455047\|2020- | ···················· | ···················· | ···················· | ···················· | ···················· | ···················· | |
| C | 8633 | hCoV-19/Australia/NSW508/2020\|EPI_ISL_455048\|2020- | ···················· | ···················· | ···················· | ···················· | ···················· | ···················· | |
| C | 8634 | hCoV-19/Australia/NSW509/2020\|EPI_ISL_455049\|2020- | ···················· | ···················· | ···················· | ···················· | ···················· | ···················· | |
| C | 8635 | hCoV-19/Australia/NSW510/2020\|EPI_ISL_455050\|2020- | ···················· | ···················· | ···················· | ···················· | ···················· | ···················· | |
| C | 8636 | hCoV-19/Australia/NSW511/2020\|EPI_ISL_455051\|2020- | ···················· | ···················· | ···················· | ···················· | ···················· | ···················· | |
| C | 8637 | hCoV-19/Australia/NSW512/2020\|EPI_ISL_455052\|2020- | ···················· | ···················· | ···················· | ···················· | ···················· | ···················· | |
| C | 8638 | hCoV-19/Australia/NSW513/2020\|EPI_ISL_455053\|2020- | ···················· | ···················· | ···················· | ···················· | ···················· | ···················· | |
| C | 8639 | hCoV-19/Australia/NSW514/2020\|EPI_ISL_455054\|2020- | ···················· | ···················· | ···················· | ···················· | ···················· | ···················· | |
| C | 8640 | hCoV-19/Australia/NSW515/2020\|EPI_ISL_455055\|2020- | ···················· | ···················· | ···················· | ···················· | ···················· | ···················· | |
| C | 8641 | hCoV-19/Australia/NSW516/2020\|EPI_ISL_455056\|2020- | ···················· | ···················· | ···················· | ···················· | ···················· | ···················· | |
| C | 8642 | hCoV-19/Australia/NSW517/2020\|EPI_ISL_455057\|2020- | ···················· | ···················· | ···················· | ···················· | ···················· | ···················· | |
| C | 8643 | hCoV-19/Australia/NSW518/2020\|EPI_ISL_455058\|2020- | ···················· | ···················· | ···················· | ···················· | ···················· | ···················· | |
| C | 8644 | hCoV-19/Australia/NSW519/2020\|EPI_ISL_455059\|2020- | ···················· | ···················· | ···················· | ···················· | ···················· | ···················· | |
| C | 8645 | hCoV-19/New Zealand/20VR2073/2020\|EPI_ISL_456360\|2 | ···················· | ···················· | ···················· | ···················· | ···················· | ···················· | |
| C | 8646 | hCoV-19/New Zealand/20VR2075/2020\|EPI_ISL_456362\|2 | ···················· | ···················· | ···················· | ···················· | ···················· | ···················· | |
| C | 8647 | hCoV-19/New Zealand/20VR2076/2020\|EPI_ISL_456363\|2 | ···················· | ···················· | ···················· | ···················· | ···················· | ···················· | |
| C | 8648 | hCoV-19/New Zealand/20VR2077/2020\|EPI_ISL_456364\|2 | ···················· | ···················· | ···················· | ···················· | ···················· | ···················· | |
| C | 8649 | hCoV-19/New Zealand/20VR2078/2020\|EPI_ISL_456365\|2 | ···················· | ···················· | ···················· | ···················· | ···················· | ···················· | |
| C | 8650 | hCoV-19/New Zealand/20VR2079/2020\|EPI_ISL_456366\|2 | ···················· | ···················· | ···················· | ···················· | ···················· | ···················· | |
| C | 8651 | hCoV-19/New Zealand/20VR2081/2020\|EPI_ISL_456367\|2 | ···················· | ···················· | ···················· | ···················· | ···················· | ···················· | |
| C | 8652 | hCoV-19/New Zealand/20VR2085/2020\|EPI_ISL_456369\|2 | ···················· | ···················· | ···················· | ···················· | ···················· | ···················· | |
| C | 8653 | hCoV-19/New Zealand/20VR2086/2020\|EPI_ISL_456370\|2 | ···················· | ···················· | ···················· | ···················· | ···················· | ···················· | |
| C | 8654 | hCoV-19/New Zealand/20VR2088/2020\|EPI_ISL_456371\|2 | ···················· | ···················· | ···················· | ···················· | ···················· | ···················· | |
| C | 8655 | hCoV-19/New Zealand/20VR2093/2020\|EPI_ISL_456374\|2 | ···················· | ···················· | ···················· | ···················· | ···················· | ···················· | |
| C | 8656 | hCoV-19/New Zealand/20VR2094/2020\|EPI_ISL_456375\|2 | ···················· | ···················· | ···················· | ···················· | ···················· | ···················· | |
| C | 8657 | hCoV-19/New Zealand/20VR2097/2020\|EPI_ISL_456376\|2 | ···················· | ···················· | ···················· | ···················· | ···················· | ···················· | |
| C | 8658 | hCoV-19/New Zealand/20VR2108/2020\|EPI_ISL_456377\|2 | ···················· | ···················· | ···················· | ···················· | ···················· | ···················· | |
| C | 8659 | hCoV-19/New Zealand/20VR2109/2020\|EPI_ISL_456378\|2 | ···················· | ···················· | ···················· | ···················· | ···················· | ···················· | |
| C | 8660 | hCoV-19/New Zealand/20VR2475/2020\|EPI_ISL_456379\|2 | ···················· | ···················· | ···················· | ···················· | ···················· | ···················· | |
| C | 8661 | hCoV-19/New Zealand/20VR2485/2020\|EPI_ISL_456380\|2 | ···················· | ···················· | ···················· | ···················· | ···················· | ···················· | |
| C | 8662 | hCoV-19/New Zealand/20VR2495/2020\|EPI_ISL_456381\|2 | ···················· | ···················· | ···················· | ···················· | ···················· | ···················· | |
| C | 8663 | hCoV-19/New Zealand/20VR2522/2020\|EPI_ISL_456382\|2 | ···················· | ···················· | ···················· | ···················· | ···················· | ···················· | |
| C | 8664 | hCoV-19/New Zealand/20VR2539/2020\|EPI_ISL_456383\|2 | ···················· | ···················· | ···················· | ···················· | ···················· | ···················· | |
| C | 8665 | hCoV-19/New Zealand/20VR2541/2020\|EPI_ISL_456384\|2 | ···················· | ···················· | ···················· | ···················· | ···················· | ···················· | |
| C | 8666 | hCoV-19/New Zealand/20VR2554/2020\|EPI_ISL_456385\|2 | ···················· | ···················· | ···················· | ···················· | ···················· | ···················· | |
| C | 8667 | hCoV-19/New Zealand/20VR2659/2020\|EPI_ISL_456388\|2 | ···················· | ···················· | ···················· | ···················· | ···················· | ···················· | |
| C | 8668 | hCoV-19/England/NORW-EAFA5/2020\|EPI_ISL_457401\|202 | ···················· | ···················· | ···················· | ···················· | ···················· | ···················· | |
| C | 8669 | hCoV-19/England/NORW-EAFC3/2020\|EPI_ISL_457402\|202 | ···················· | ···················· | ···················· | ···················· | ···················· | ···················· | |
| C | 8670 | hCoV-19/England/NORW-EAFD2/2020\|EPI_ISL_457403\|202 | ···················· | ···················· | ···················· | ···················· | ···················· | ···················· | |
| C | 8671 | hCoV-19/Guangzhou/GZMU0078/2020\|EPI_ISL_457690\|202 | ···················· | ···················· | ···················· | ···················· | ···················· | ···················· | |
| C | 8672 | hCoV-19/Guangzhou/GZMU0079/2020\|EPI_ISL_457691\|202 | ···················· | ·A·················· | ···················· | ···················· | ···················· | ···················· | |
| C | 8673 | hCoV-19/Guangzhou/GZMU0081/2020\|EPI_ISL_457692\|202 | ···················· | ···················· | ···················· | ···················· | ···················· | ···················· | |
| C | 8674 | hCoV-19/Guangzhou/GZMU0083/2020\|EPI_ISL_457693\|202 | ···················· | ···················· | ···················· | ···················· | ···················· | ···················· | |
| C | 8675 | hCoV-19/Guangzhou/GZMU0092/2020\|EPI_ISL_457694\|202 | ···················· | ·A·················· | ···················· | ···················· | ···················· | ···················· | |
| C | 8676 | hCoV-19/Guangzhou/GZMU0095/2020\|EPI_ISL_457695\|202 | ···················· | ···················· | ···················· | ···················· | ···················· | ···················· | |
| C | 8677 | hCoV-19/Guangzhou/GZMU0099/2020\|EPI_ISL_457696\|202 | ···················· | ···················· | ···················· | ···················· | ···················· | ···················· | |
| C | 8678 | hCoV-19/Guangzhou/GZMU0101/2020\|EPI_ISL_457697\|202 | ···················· | ···················· | ···················· | ···················· | ···················· | ···················· | |
| C | 8679 | hCoV-19/Guangzhou/GZMU0117/2020\|EPI_ISL_457698\|202 | ···················· | ···················· | ···················· | ···················· | ···················· | ···················· | |
| C | 8680 | hCoV-19/Italy/EMR-AMVRC-206/2020\|EPI_ISL_457699\|20 | ···················· | ···················· | ···················· | ···················· | ···················· | ···················· | |
| C | 8681 | hCoV-19/England/NORT-288F5C/2020\|EPI_ISL_458700\|20 | ···················· | ···················· | ···················· | ···················· | ···················· | ···················· | |
| C | 8682 | hCoV-19/England/NORT-2893C8/2020\|EPI_ISL_458701\|20 | ···················· | ···················· | ···················· | ···················· | ···················· | ···················· | |
| C | 8683 | hCoV-19/England/NORT-2898F6/2020\|EPI_ISL_458702\|20 | ···················· | ···················· | ···················· | ···················· | ···················· | ···················· | |
| C | 8684 | hCoV-19/England/NORT-289340/2020\|EPI_ISL_458703\|20 | ···················· | ···················· | ···················· | ···················· | ···················· | ···················· | |
| C | 8685 | hCoV-19/England/NORT-289665/2020\|EPI_ISL_458706\|20 | ···················· | ···················· | ···················· | ···················· | ···················· | ···················· | |
| C | 8686 | hCoV-19/England/NORT-2891EC/2020\|EPI_ISL_458707\|20 | ···················· | ···················A | ···················· | ···················· | ···················· | ···················· | 1 |
| C | 8687 | hCoV-19/England/NORT-289085/2020\|EPI_ISL_458708\|20 | ···················· | ···················· | ···················· | ···················· | ···················· | ···················· | |
| C | 8688 | hCoV-19/England/NORT-2890D0/2020\|EPI_ISL_458709\|20 | ···················· | ···················· | ···················· | ···················· | ···················· | ···················· | |
| C | 8689 | hCoV-19/England/NORT-289331/2020\|EPI_ISL_458710\|20 | ···················· | ···················· | ···················· | ···················· | ···················· | ···················· | |
| C | 8690 | hCoV-19/England/NORT-28939B/2020\|EPI_ISL_458711\|20 | ···················· | ···················· | ···················· | ···················· | ···················· | ···················· | |
| C | 8691 | hCoV-19/England/NORT-2892F8/2020\|EPI_ISL_458713\|20 | ···················· | ···················· | ···················· | ···················· | ···················· | ···················· | |
| C | 8692 | hCoV-19/England/NORT-2897DB/2020\|EPI_ISL_458714\|20 | ···················· | ···················· | ···················· | ···················· | ···················· | ···················· | |
| C | 8693 | hCoV-19/England/NORT-2892AD/2020\|EPI_ISL_458715\|20 | ···················· | ···················A | ···················· | ···················· | ···················· | ···················· | 1 |
| C | 8694 | hCoV-19/England/NORT-28942F/2020\|EPI_ISL_458717\|20 | ···················· | ·A·················· | ···················· | ···················· | ···················· | ···················· | |
| C | 8695 | hCoV-19/England/NORT-28954A/2020\|EPI_ISL_458718\|20 | ···················· | ···················· | ···················· | ···················· | ···················· | ···················· | |
| C | 8696 | hCoV-19/England/BRIS-12911C/2020\|EPI_ISL_458721\|20 | ···················· | ···················· | ···················· | ···················· | ···················· | ···················· | |
| C | 8697 | hCoV-19/England/BRIS-129422/2020\|EPI_ISL_458722\|20 | ···················· | ···················· | ···················· | ···················· | ···················· | ···················· | |
| C | 8698 | hCoV-19/England/BRIS-128F22/2020\|EPI_ISL_458723\|20 | ···················· | ···················· | ···················· | ···················· | ···················· | ···················· | |
| C | 8699 | hCoV-19/England/BRIS-128AB8/2020\|EPI_ISL_458725\|20 | ···················· | ·············A······ | ···················· | ···················· | ···················· | ···················· | |
| C | 8700 | hCoV-19/England/BRIS-1291FE/2020\|EPI_ISL_458726\|20 | ···················· | ···················· | ···················· | ···················· | ···················· | ···················· | |
| C | 8701 | hCoV-19/England/BRIS-128881/2020\|EPI_ISL_458727\|20 | ···················· | ···················· | ···················· | ···················· | ···················· | ···················· | |
| C | 8702 | hCoV-19/England/BRIS-1289BB/2020\|EPI_ISL_458728\|20 | ···················· | ···················· | ···················· | ···················· | ···················· | ···················· | |

C 8703 hCoV-19/England/BRIS-128B2E/2020|EPI_ISL_458732|20 ···················· ···················· ···················· ···················· ···················· ····················
C 8704 hCoV-19/England/BRIS-129149/2020|EPI_ISL_458733|20 ···················· ····A··············· ···················· ···················· ···················· ····················
C 8705 hCoV-19/England/CAMB-1AD14D/2020|EPI_ISL_447970|20 ···················· ···················· ···················· ···················· ···················· ····················
C 8706 hCoV-19/England/CAMB-1AD16B/2020|EPI_ISL_447971|20 ···················· ···················· ···················· ···················· ···················· ····················
C 8707 hCoV-19/England/CAMB-1AD17A/2020|EPI_ISL_447972|20 ···················· ···················· ···················· ···················· ···················· ····················
C 8708 hCoV-19/England/CAMB-1AD189/2020|EPI_ISL_447973|20 ···················· ···················· ···················· ···················· ···················· ····················
C 8709 hCoV-19/England/CAMB-1AD1C5/2020|EPI_ISL_447976|20 ···················· ···················· ···················· ···················· ···················· ····················
C 8710 hCoV-19/England/CAMB-1AD2D1/2020|EPI_ISL_447988|20 ···················· ·A·················· ···················· ···················· ···················· ····················
C 8711 hCoV-19/England/CAMB-1AD2E0/2020|EPI_ISL_447989|20 ···················· ···················· ···················· ···················· ···················· ····················
C 8712 hCoV-19/England/CAMB-1AD602/2020|EPI_ISL_447990|20 ···················· ···················· ···················· ···················· ···················· ····················
C 8713 hCoV-19/England/CAMB-1AD6A8/2020|EPI_ISL_447994|20 ···················· ···················· ···················· ···················· ···················· ····················
C 8714 hCoV-19/England/CAMB-1AD6B7/2020|EPI_ISL_447995|20 ···················· ···················· ···················· ···················· ···················· ····················
C 8715 hCoV-19/England/CAMB-1AD6D5/2020|EPI_ISL_447997|20 ···················· ···················· ···················· ···················· ···················· ····················
C 8716 hCoV-19/England/CAMB-1AD6F3/2020|EPI_ISL_447998|20 ···················· ···················· ···················· ···················· ···················· ····················
C 8717 hCoV-19/England/CAMB-1AD70F/2020|EPI_ISL_447999|20 ···················· ···················· ···················· ···················· ···················· ····················
C 8718 hCoV-19/USA/LA-EVTL061/2020|EPI_ISL_451222|2020-04 ···················· ···················· ···················· ···················· ···················· ····················
C 8719 hCoV-19/USA/LA-EVTL062/2020|EPI_ISL_451223|2020-04 ···················· ···················· ···················· ···················· ···················· ····················
C 8720 hCoV-19/USA/LA-EVTL063/2020|EPI_ISL_451224|2020-04 ···················· ···················· ···················· ···················· ···················· ····················
C 8721 hCoV-19/USA/LA-EVTL064/2020|EPI_ISL_451225|2020-04 ···················· ···················· ···················· ···················· ···················· ····················
C 8722 hCoV-19/USA/LA-EVTL065/2020|EPI_ISL_451226|2020-05 ···················· ···················· ···················· ···················· ···················· ····················
C 8723 hCoV-19/USA/LA-EVTL066/2020|EPI_ISL_451227|2020-05 ···················· ···················· ···················· ···················· ···················· ····················
C 8724 hCoV-19/USA/LA-EVTL067/2020|EPI_ISL_451228|2020-04 ···················· ···················· ···················· ···················· ···················· ····················
C 8725 hCoV-19/USA/LA-EVTL068/2020|EPI_ISL_451229|2020-04 ···················· ···················· ···················· ···················· ···················· ····················
C 8726 hCoV-19/USA/LA-EVTL069/2020|EPI_ISL_451230|2020-04 ···················· ···················· ···················· ···················· ···················· ····················
C 8727 hCoV-19/USA/LA-EVTL070/2020|EPI_ISL_451231|2020-05 ···················· ···················· ···················· ···················· ···················· ····················
C 8728 hCoV-19/USA/LA-EVTL071/2020|EPI_ISL_451232|2020-05 ···················· ···················· ···················· ···················· ···················· ····················
C 8729 hCoV-19/USA/LA-EVTL078/2020|EPI_ISL_451239|2020-05 ···················· ···················· ···················· ···················· ···················· ····················
C 8730 hCoV-19/USA/LA-EVTL079/2020|EPI_ISL_451240|2020-05 ···················· ···················· ···················· ···················· ···················· ····················
C 8731 hCoV-19/USA/LA-EVTL080/2020|EPI_ISL_451241|2020-05 ···················· ···················· ···················· ···················· ···················· ····················
C 8732 hCoV-19/USA/LA-EVTL081/2020|EPI_ISL_451242|2020-05 ···················· ···················· ···················· ···················· ···················· ····················
C 8733 hCoV-19/USA/LA-EVTL082/2020|EPI_ISL_451243|2020-05 ···················· ···················· ···················· ···················· ···················· ····················
C 8735 hCoV-19/Spain/NC-IBV-001398/2020|EPI_ISL_452503|20 ···················· ···················· ···················· ···················· ···················· ····················
C 8739 hCoV-19/Spain/NC-IBV-001393/2020|EPI_ISL_452508|20 ···················· ···················· ···················· ···················· ···················· ····················
C 8744 hCoV-19/Spain/AN-IBV-001217/2020|EPI_ISL_452556|20 ···················· ···················· ···················· ···················· ··N················· ····················
C 8745 hCoV-19/Spain/AN-IBV-001216/2020|EPI_ISL_452557|20 ···················· ···················· ···················· ···················· ···················· ····················
C 8749 hCoV-19/Spain/AN-IBV-001209/2020|EPI_ISL_452561|20 ···················· ···················· ···················· ···················· ···················· ····················
C 8751 hCoV-19/Spain/AN-IBV-001169/2020|EPI_ISL_452570|20 ···················· ···················· ···················· ···················· ···················· ····················
C 8756 hCoV-19/Portugal/PT0097/2020|EPI_ISL_453813|2020-0 ···················· ···················· ···················· ···················· ···················· ····················
C 8757 hCoV-19/Portugal/PT0098/2020|EPI_ISL_453814|2020-0 ···················· ···················· ···················· ···················· ···················· ····················
C 8758 hCoV-19/Portugal/PT0099/2020|EPI_ISL_453815|2020-0 ···················· ···················· ···················· ···················· ···················· ····················
C 8759 hCoV-19/Portugal/PT0100/2020|EPI_ISL_453816|2020-0 ···················· ···················· ···················· ···················· ···················· ····················
C 8760 hCoV-19/Portugal/PT0101/2020|EPI_ISL_453817|2020-0 ···················· ···················· ···················· ···················· ···················· ····················
C 8761 hCoV-19/Portugal/PT0102/2020|EPI_ISL_453818|2020-0 ···················· ···················· ···················· ···················· ···················· ····················
C 8762 hCoV-19/Portugal/PT0104/2020|EPI_ISL_453820|2020-0 ···················· ···················· ···················· ···················· ···················· ····················
C 8763 hCoV-19/Portugal/PT0105/2020|EPI_ISL_453821|2020-0 ···················· ···················· ···················· ···················· ···················· ····················
C 8764 hCoV-19/Portugal/PT0106/2020|EPI_ISL_453822|2020-0 ···················· ···················· ···················· ···················· ···················· ····················
C 8765 hCoV-19/Portugal/PT0107/2020|EPI_ISL_453823|2020-0 ···················· ···················· ···················· ···················· ···················· ····················
C 8766 hCoV-19/Portugal/PT0108/2020|EPI_ISL_453824|2020-0 ···················· ···················· ···················· ···················· ···················· ····················
C 8767 hCoV-19/Portugal/PT0109/2020|EPI_ISL_453825|2020-0 ···················· ···················· ···················· ···················· ···················· ····················
C 8768 hCoV-19/Portugal/PT0110/2020|EPI_ISL_453826|2020-0 ···················· ···················· ···················· ···················· ···················· ····················
C 8769 hCoV-19/Portugal/PT0111/2020|EPI_ISL_453827|2020-0 ···················· ···················· ···················· ···················· ···················· ····················
C 8770 hCoV-19/Portugal/PT0112/2020|EPI_ISL_453828|2020-0 ···················· ···················· ···················· ···················· ···················· ····················
C 8771 hCoV-19/Portugal/PT0113/2020|EPI_ISL_453829|2020-0 ···················· ···················· ···················· ···················· ···················· ····················
C 8772 hCoV-19/Portugal/PT0114/2020|EPI_ISL_453830|2020-0 ···················· ···················· ···················· ···················· ···················· ····················
C 8773 hCoV-19/Portugal/PT0116/2020|EPI_ISL_453832|2020-0 ···················· ···················· ···················· ···················· ···················· ····················
C 8774 hCoV-19/Portugal/PT0117/2020|EPI_ISL_453833|2020-0 ···················· ···················· ···················· ···················· ···················· ····················
C 8775 hCoV-19/Portugal/PT0118/2020|EPI_ISL_453834|2020-0 ···················· ···················· ···················· ···················· ···················· ····················
C 8776 hCoV-19/Portugal/PT0119/2020|EPI_ISL_453835|2020-0 ···················· ···················· ···················· ···················· ···················· ····················
C 8777 hCoV-19/Portugal/PT0120/2020|EPI_ISL_453836|2020-0 ···················· ···················· ···················· ···················· ···················· ····················
C 8778 hCoV-19/Portugal/PT0123/2020|EPI_ISL_453839|2020-0 ···················· ···················· ···················· ···················· ···················· ····················
C 8779 hCoV-19/Portugal/PT0168/2020|EPI_ISL_453884|2020-0 ···················· ···················· ···················· ···················· ···················· ····················
C 8780 hCoV-19/Portugal/PT0169/2020|EPI_ISL_453885|2020-0 ···················· ···················· ···················· ···················· ···················· ····················
C 8781 hCoV-19/Portugal/PT0171/2020|EPI_ISL_453887|2020-0 ···················· ···················· ···················· ···················· ···················· ····················
C 8782 hCoV-19/Portugal/PT0173/2020|EPI_ISL_453889|2020-0 ···················· ···················· ···················· ···················· ···················· ····················
C 8783 hCoV-19/Portugal/PT0174/2020|EPI_ISL_453890|2020-0 ···················· ···················· ···················· ···················· ···················· ····················
C 8784 hCoV-19/Portugal/PT0175/2020|EPI_ISL_453891|2020-0 ···················· ···················· ···················· ···················· ···················· ····················
C 8785 hCoV-19/Portugal/PT0176/2020|EPI_ISL_453892|2020-0 ···················· ···················· ···················· ···················· ···················· ····················
C 8786 hCoV-19/Portugal/PT0177/2020|EPI_ISL_453893|2020-0 ···················· ···················· ···················· ···················· ···················· ····················
C 8787 hCoV-19/Portugal/PT0178/2020|EPI_ISL_453894|2020-0 ···················· ···················· ···················· ···················· ···················· ····················
C 8788 hCoV-19/USA/NY-NYUMC804/2020|EPI_ISL_456040|2020-0 ···················· ···················· ···················· ···················· ···················· ····················
C 8789 hCoV-19/USA/NY-NYUMC805/2020|EPI_ISL_456041|2020-0 ···················· ···················· ···················· ···················· ···················· ····················
C 8790 hCoV-19/USA/NY-NYUMC806/2020|EPI_ISL_456042|2020-0 ···················· ···················· ···················· ···················· ···················· ····················
C 8791 hCoV-19/USA/NY-NYUMC807/2020|EPI_ISL_456043|2020-0 ···················· ···················· ···················· ···················· ···················· ····················
C 8792 hCoV-19/USA/NY-NYUMC808/2020|EPI_ISL_456044|2020-0 ···················· ···················· ···················· ···················· ···················· ····················
C 8793 hCoV-19/USA/NY-NYUMC809/2020|EPI_ISL_456045|2020-0 ···················· ···················· ···················· ···················· ···················· ····················
C 8794 hCoV-19/USA/NY-NYUMC810/2020|EPI_ISL_456046|2020-0 ···················· ···················· ···················· ···················· ···················· ····················
C 8795 hCoV-19/USA/NY-NYUMC811/2020|EPI_ISL_456047|2020-0 ···················· ···················· ···················· ···················· ···················· ····················
C 8796 hCoV-19/USA/NY-NYUMC813/2020|EPI_ISL_456049|2020-0 ···················· ···················· ···················· ···················· ···················· ····················

| | | | | | | | |
|---|---|---|---|---|---|---|---|
| C | 8797 hCoV-19/USA/NY-NYUMC814/2020\|EPI_ISL_456050\|2020-0 | ···················· | ···················· | ····················· | ···················· | ···················· | ···················· |
| C | 8798 hCoV-19/USA/NJ-NYUMC815/2020\|EPI_ISL_456051\|2020-0 | ···················· | ···················· | ····················· | ···················· | ···················· | ···················· |
| C | 8799 hCoV-19/USA/NY-NYUMC816/2020\|EPI_ISL_456052\|2020-0 | ···················· | ···················· | ····················· | ···················· | ···················· | ···················· |
| C | 8800 hCoV-19/USA/NY-NYUMC817/2020\|EPI_ISL_456053\|2020-0 | ···················· | ···················· | ····················· | ···················· | ···················· | ···················· |
| C | 8801 hCoV-19/USA/NY-NYUMC818/2020\|EPI_ISL_456054\|2020-0 | ···················· | ···················· | ····················· | ···················· | ···················· | ···················· |
| C | 8802 hCoV-19/USA/NY-NYUMC819/2020\|EPI_ISL_456055\|2020-0 | ···················· | ···················· | ····················· | ···················· | ···················· | ···················· |
| C | 8803 hCoV-19/USA/NY-NYUMC820/2020\|EPI_ISL_456056\|2020-0 | ···················· | ···················· | ····················· | ···················· | ···················· | ···················· |
| C | 8804 hCoV-19/USA/NY-NYUMC822/2020\|EPI_ISL_456058\|2020-0 | ···················· | ···················· | ····················· | ···················· | ···················· | ···················· |
| C | 8805 hCoV-19/USA/NY-NYUMC824/2020\|EPI_ISL_456060\|2020-0 | ···················· | ···················· | ····················· | ···················· | ···················· | ···················· |
| C | 8806 hCoV-19/USA/NY-NYUMC825/2020\|EPI_ISL_456061\|2020-0 | ···················· | ···················· | ····················· | ···················· | ···················· | ···················· |
| C | 8807 hCoV-19/USA/NY-NYUMC826/2020\|EPI_ISL_456062\|2020-0 | ···················· | ···················· | ····················· | ···················· | ···················· | ···················· |
| C | 8808 hCoV-19/USA/NY-NYUMC827/2020\|EPI_ISL_456063\|2020-0 | ···················· | ···················· | ····················· | ···················· | ···················· | ···················· |
| C | 8809 hCoV-19/USA/NY-NYUMC829/2020\|EPI_ISL_456065\|2020-0 | ···················· | ···················· | ····················· | ···················· | ···················· | ···················· |
| C | 8810 hCoV-19/USA/NY-NYUMC791/2020\|EPI_ISL_456066\|2020-0 | ···················· | ···················· | ····················· | ···················· | ···················· | ···················· |
| C | 8811 hCoV-19/Brazil/RJ-899/2020\|EPI_ISL_456071\|2020-03- | ···················· | ···················· | ····················· | ···················· | ···················· | ···················· |
| C | 8812 hCoV-19/Brazil/RJ-1056/2020\|EPI_ISL_456072\|2020-04 | ···················· | ···················· | ····················· | ···················· | ···················· | ···················· |
| C | 8813 hCoV-19/Brazil/RJ-1058/2020\|EPI_ISL_456073\|2020-04 | ···················· | ···················· | ····················· | ···················· | ···················· | ···················· |
| C | 8814 hCoV-19/Brazil/RJ-1065/2020\|EPI_ISL_456074\|2020-04 | ···················· | ···················· | ····················· | ···················· | ···················· | ···················· |
| C | 8815 hCoV-19/Brazil/RJ-1100/2020\|EPI_ISL_456075\|2020-04 | ···················· | ···················· | ····················· | ···················· | ···················· | ···················· |
| C | 8816 hCoV-19/Brazil/RJ-1111/2020\|EPI_ISL_456076\|2020-03 | ···················· | ···················· | ····················· | ···················· | ···················· | ···················· |
| C | 8817 hCoV-19/Brazil/RJ-1119/2020\|EPI_ISL_456077\|2020-03 | ···················· | ···················· | ····················· | ···················· | ···················· | ···················· |
| C | 8818 hCoV-19/Brazil/RJ-1402/2020\|EPI_ISL_456079\|2020-04 | ···················· | ···················· | ····················· | ···················· | ···················· | ···················· |
| C | 8819 hCoV-19/Brazil/RJ-1464/2020\|EPI_ISL_456080\|2020-04 | ···················· | ···················· | ····················· | ···················· | ···················· | ···················· |
| C | 8820 hCoV-19/Brazil/RJ-1466/2020\|EPI_ISL_456081\|2020-04 | ···················· | ···················· | ····················· | ···················· | ···················· | ···················· |
| C | 8821 hCoV-19/Brazil/RJ-1600/2020\|EPI_ISL_456082\|2020-04 | ···················· | ···················· | ····················· | ···················· | ···················· | ···················· |
| C | 8822 hCoV-19/Brazil/RJ-1627/2020\|EPI_ISL_456083\|2020-04 | ···················· | ···················· | ····················· | ···················· | ···················· | ···················· |
| C | 8823 hCoV-19/Brazil/RJ-1690/2020\|EPI_ISL_456084\|2020-04 | ···················· | ···················· | ····················· | ···················· | ···················· | ···················· |
| C | 8824 hCoV-19/Brazil/RJ-1691/2020\|EPI_ISL_456085\|2020-04 | ···················· | ···················· | ····················· | ···················· | ···················· | ···················· |
| C | 8825 hCoV-19/Brazil/RJ-1701/2020\|EPI_ISL_456086\|2020-04 | ···················· | ···················· | ····················· | ···················· | ···················· | ···················· |
| C | 8826 hCoV-19/Brazil/RJ-1702/2020\|EPI_ISL_456087\|2020-04 | ···················· | ···················· | ····················· | ···················· | ···················· | ···················· |
| C | 8827 hCoV-19/Brazil/RJ-1719/2020\|EPI_ISL_456088\|2020-04 | ···················· | ···················· | ····················· | ···················· | ···················· | ···················· |
| C | 8828 hCoV-19/Brazil/RJ-1901/2020\|EPI_ISL_456089\|2020-04 | ···················· | ···················· | ····················· | ···················· | ···················· | ···················· |
| C | 8829 hCoV-19/Brazil/RJ-1902/2020\|EPI_ISL_456090\|2020-04 | ···················· | ···················· | ····················· | ···················· | ···················· | ···················· |
| C | 8830 hCoV-19/Brazil/RJ-1921/2020\|EPI_ISL_456091\|2020-04 | ···················· | ···················· | ····················· | ···················· | ···················· | ···················· |
| C | 8831 hCoV-19/Brazil/RJ-1923/2020\|EPI_ISL_456092\|2020-04 | ···················· | ···················· | ····················· | ···················· | ···················· | ···················· |
| C | 8832 hCoV-19/Brazil/RJ-1927/2020\|EPI_ISL_456093\|2020-04 | ···················· | ···················· | ····················· | ···················· | ···················· | ···················· |
| C | 8833 hCoV-19/Brazil/RJ-1943/2020\|EPI_ISL_456094\|2020-04 | ···················· | ···················· | ····················· | ···················· | ···················· | ···················· |
| C | 8834 hCoV-19/Brazil/RJ-1948/2020\|EPI_ISL_456095\|2020-04 | ···················· | ···················· | ····················· | ···················· | ···················· | ···················· |
| C | 8835 hCoV-19/Brazil/RJ-1952/2020\|EPI_ISL_456096\|2020-04 | ··············G····· | ···················· | ····················· | ···················· | ···················· | ···················· |
| C | 8836 hCoV-19/Brazil/RJ-1966/2020\|EPI_ISL_456097\|2020-04 | ···················· | ···················· | ····················· | ···················· | ···················· | ···················· |
| C | 8837 hCoV-19/Brazil/RJ-2000/2020\|EPI_ISL_456098\|2020-04 | ···················· | ···················· | ····················· | ···················· | ···················· | ···················· |
| C | 8838 hCoV-19/Brazil/RJ-2007/2020\|EPI_ISL_456099\|2020-04 | ···················· | ···················· | ····················· | ···················· | ···················· | ···················· |
| C | 8839 hCoV-19/England/NORT-284EAE/2020\|EPI_ISL_457303\|20 | ···················· | ····A··············· | ····················· | ···················· | ···················· | ···················· |
| C | 8840 hCoV-19/England/NORT-28A558/2020\|EPI_ISL_457305\|20 | ···················· | ···················· | ····················· | ···················· | ···················· | ···················· |
| C | 8841 hCoV-19/England/NORT-28B302/2020\|EPI_ISL_457311\|20 | ···················· | ···················· | ····················· | ···················· | ···················· | ···················· |
| C | 8842 hCoV-19/England/NORT-28B38A/2020\|EPI_ISL_457312\|20 | ···················· | ···················· | ····················· | ···················· | ···················· | ···················· |
| C | 8843 hCoV-19/England/NORT-28BF1D/2020\|EPI_ISL_457316\|20 | ···················· | ···················· | ····················· | ···················· | ···················· | ···················· |
| C | 8844 hCoV-19/England/NORT-292238/2020\|EPI_ISL_457319\|20 | ···················· | ···················· | ····················· | ···················· | ···················· | ···················· |
| C | 8845 hCoV-19/England/NORT-292317/2020\|EPI_ISL_457320\|20 | ···················· | ···················· | ····················· | ···················· | ···················· | ···················· |
| C | 8846 hCoV-19/England/NORT-292739/2020\|EPI_ISL_457325\|20 | ···················· | ···················· | ····················· | ···················· | ···················· | ···················· |
| C | 8847 hCoV-19/England/NORW-E752E/2020\|EPI_ISL_457326\|202 | ···················· | ···················· | ····················· | ···················· | ···················· | ···················· |
| C | 8848 hCoV-19/England/NORW-EACF9/2020\|EPI_ISL_457370\|202 | ···················· | ···················· | ····················· | ···················· | ···················· | ···················· |
| C | 8849 hCoV-19/England/NORW-EAD32/2020\|EPI_ISL_457371\|202 | ···················· | ···················· | ····················· | ···················· | ···················· | ···················· |
| C | 8850 hCoV-19/England/NORW-EAD7E/2020\|EPI_ISL_457373\|202 | ···················· | ···················· | ····················· | ···················· | ···················· | ···················· |
| C | 8851 hCoV-19/England/NORW-EAD80/2020\|EPI_ISL_457374\|202 | ···················· | ···················· | ····················· | ···················· | ···················· | ···················· |
| C | 8852 hCoV-19/England/NORW-EAD9C/2020\|EPI_ISL_457375\|202 | ···················· | ···················· | ····················· | ···················· | ···················· | ···················· |
| C | 8853 hCoV-19/England/NORW-EADBA/2020\|EPI_ISL_457377\|202 | ···················· | ···················· | ····················· | ···················· | ···················· | ···················· |
| C | 8854 hCoV-19/England/NORW-EADD8/2020\|EPI_ISL_457378\|202 | ···················· | ···················· | ····················· | ···················· | ···················· | ···················· |
| C | 8855 hCoV-19/England/NORW-EADF6/2020\|EPI_ISL_457379\|202 | ···················· | ···················· | ····················· | ···················· | ···················· | ···················· |
| C | 8856 hCoV-19/England/NORW-EAE02/2020\|EPI_ISL_457380\|202 | ···················· | ···················· | ····················· | ···················· | ···················· | ···················· |
| C | 8857 hCoV-19/England/NORW-EAE11/2020\|EPI_ISL_457381\|202 | ···················· | ···················· | ····················· | ···················· | ···················· | ···················· |
| C | 8858 hCoV-19/England/NORW-EAE20/2020\|EPI_ISL_457382\|202 | ···················· | ···················· | ····················· | ···················· | ···················· | ···················· |
| C | 8859 hCoV-19/England/NORW-EAE3F/2020\|EPI_ISL_457383\|202 | ···················· | ···················· | ····················· | ···················· | ···················· | ···················· |
| C | 8860 hCoV-19/England/NORW-EAE6C/2020\|EPI_ISL_457385\|202 | ···················· | ···················· | ····················· | ···················· | ···················· | ···················· |
| C | 8861 hCoV-19/England/NORW-EAE7B/2020\|EPI_ISL_457386\|202 | ···················· | ···················· | ····················· | ···················· | ···················· | ···················· |
| C | 8862 hCoV-19/England/NORW-EAE8A/2020\|EPI_ISL_457387\|202 | ···················· | ···················· | ····················· | ···················· | ···················· | ···················· |
| C | 8863 hCoV-19/England/NORW-EAE99/2020\|EPI_ISL_457388\|202 | ···················· | ···················· | ····················· | ···················· | ···················· | ···················· |
| C | 8864 hCoV-19/England/NORW-EAEA8/2020\|EPI_ISL_457389\|202 | ···················· | ···················· | ····················· | ···················· | ···················· | ···················· |
| C | 8865 hCoV-19/England/NORW-EAEB7/2020\|EPI_ISL_457390\|202 | ···················· | ···················· | ····················· | ···················· | ···················· | ···················· |
| C | 8866 hCoV-19/England/NORW-EAED5/2020\|EPI_ISL_457391\|202 | ···················· | ···················· | ····················· | ···················· | ···················· | ···················· |
| C | 8867 hCoV-19/England/NORW-EAEE4/2020\|EPI_ISL_457392\|202 | ···················· | ···················· | ····················· | ···················· | ···················· | ···················· |
| C | 8868 hCoV-19/England/NORW-EAF3C/2020\|EPI_ISL_457394\|202 | ···················· | ···················· | ····················· | ···················· | ···················· | ···················· |
| C | 8869 hCoV-19/England/NORW-EAF4B/2020\|EPI_ISL_457395\|202 | ···················· | ···················· | ····················· | ···················· | ···················· | ···················· |
| C | 8870 hCoV-19/England/NORW-EAF69/2020\|EPI_ISL_457397\|202 | ···················· | ···················· | ····················· | ···················· | ···················· | ···················· |
| C | 8871 hCoV-19/England/NORT-288D61/2020\|EPI_ISL_458625\|20 | ···················· | ···················· | ····················· | ···················· | ···················· | ···················· |
| C | 8872 hCoV-19/England/NORT-2894B6/2020\|EPI_ISL_458626\|20 | ···················· | ···················· | ····················· | ···················· | ···················· | ···················· |
| C | 8873 hCoV-19/England/NORT-288FF2/2020\|EPI_ISL_458628\|20 | ···················· | ···················· | ····················· | ···················· | ···················· | ···················· |
| C | 8874 hCoV-19/England/NORT-288DCB/2020\|EPI_ISL_458631\|20 | ···················· | ···················· | ····················· | ···················· | ···················· | ···················· |

| | | | | | | | |
|---|---|---|---|---|---|---|---|
| C | 8875 hCoV-19/England/NORT-2895B3/2020\|EPI_ISL_458632\|20 | ···················· | ···················· | ···················· | ···················· | ···················· | ···················· |
| C | 8876 hCoV-19/England/NORT-289595/2020\|EPI_ISL_458633\|20 | ···················· | ···················· | ···················· | ···················· | ···················· | ···················· |
| C | 8877 hCoV-19/England/NORT-289717/2020\|EPI_ISL_458635\|20 | ···················· | ···················· | ···················· | ···················· | ···················· | ···················· |
| C | 8878 hCoV-19/England/NORT-28903A/2020\|EPI_ISL_458636\|20 | ···················· | ···················· | ···················· | ···················· | ···················· | ···················· |
| C | 8879 hCoV-19/England/NORT-28988D/2020\|EPI_ISL_458637\|20 | ···················· | ···················· | ···················· | ···················· | ···················· | ···················· |
| C | 8880 hCoV-19/England/NORT-288FE3/2020\|EPI_ISL_458638\|20 | ···················· | ···················· | ···················· | ···················· | ···················· | ···················· |
| C | 8881 hCoV-19/England/NORT-28928F/2020\|EPI_ISL_458640\|20 | ···················· | ···················· | ···················· | ···················· | ···················· | ···················· |
| C | 8882 hCoV-19/England/NORT-289076/2020\|EPI_ISL_458641\|20 | ···················· | ···················· | ···················· | ···················· | ···················· | ···················· |
| C | 8883 hCoV-19/England/NORT-288E40/2020\|EPI_ISL_458642\|20 | ···················· | ···················· | ···················· | ···················· | ···················· | ···················· |
| C | 8884 hCoV-19/England/NORT-289771/2020\|EPI_ISL_458643\|20 | ···················· | ···················· | ···················· | ···················· | ···················· | ···················· |
| C | 8885 hCoV-19/England/NORT-28929E/2020\|EPI_ISL_458644\|20 | ···················· | ···················· | ···················· | ···················· | ···················· | ···················· |
| C | 8886 hCoV-19/England/NORT-2894A7/2020\|EPI_ISL_458645\|20 | ···················· | ···················· | ···················· | ···················· | ···················· | ···················· |
| C | 8887 hCoV-19/England/NORT-289559/2020\|EPI_ISL_458646\|20 | ···················· | ···················· | ···················· | ···················· | ···················· | ···················· |
| C | 8888 hCoV-19/England/NORT-289726/2020\|EPI_ISL_458647\|20 | ···················· | ···················· | ···················· | ···················· | ···················· | ···················· |
| C | 8889 hCoV-19/England/NORT-2896A1/2020\|EPI_ISL_458648\|20 | ···················· | ···················· | ···················· | ···················· | ···················· | ···················· |
| C | 8890 hCoV-19/England/NORT-2896CF/2020\|EPI_ISL_458649\|20 | ···················· | ·············A······ | ···················· | ···················· | ···················· | ···················· |
| C | 8891 hCoV-19/England/NORT-288E7D/2020\|EPI_ISL_458650\|20 | ···················· | ···················· | ···················· | ···················· | ···················· | ···················· |
| C | 8892 hCoV-19/England/NORT-289049/2020\|EPI_ISL_458651\|20 | ···················· | ···················· | ···················· | ···················· | ···················· | ···················· |
| C | 8893 hCoV-19/England/NORT-289216/2020\|EPI_ISL_458652\|20 | ···················· | ···················· | ···················· | ···················· | ···················· | ···················· |
| C | 8894 hCoV-19/England/NORT-288F7A/2020\|EPI_ISL_458653\|20 | ···················· | ···················· | ···················· | ···················· | ···················· | ···················· |
| C | 8895 hCoV-19/England/NORT-288E22/2020\|EPI_ISL_458654\|20 | ···················· | ···················· | ···················· | ···················· | ···················· | ···················· |
| C | 8896 hCoV-19/England/NORT-289823/2020\|EPI_ISL_458656\|20 | ···················· | ···················· | ···················· | ···················· | ···················· | ···················· |
| C | 8897 hCoV-19/England/NORT-289586/2020\|EPI_ISL_458657\|20 | ···················· | ···················· | ···················· | ···················· | ···················· | ···················· |
| C | 8898 hCoV-19/Malaysia/IMR_WC10180/2020\|EPI_ISL_459957\|2 | ···················· | ···················· | ···················· | ···················· | ···················· | ···················· |
| C | 8899 hCoV-19/England/20104003104/2020\|EPI_ISL_459958\|20 | ···················· | ···················· | ···················· | ···················· | ···················· | ···················· |
| C | 8900 hCoV-19/England/201060022/2020\|EPI_ISL_459960\|2020 | ···················· | ···················· | ···················· | ···················· | ··N················· | ···················· |
| C | 8901 hCoV-19/England/201060024/2020\|EPI_ISL_459961\|2020 | ···················· | ···················· | ···················· | ···················· | ···················· | ···················· |
| C | 8902 hCoV-19/Germany/MV-Cento-70001814/2020\|EPI_ISL_459 | ···················· | ···················· | ···················· | ···················· | ···················· | ···················· |
| C | 8903 hCoV-19/Germany/MV-Cento-70004280/2020\|EPI_ISL_459 | ···················· | ···················· | ···················· | ···················· | ···················· | ···················· |
| C | 8904 hCoV-19/Germany/MV-Cento-70002372/2020\|EPI_ISL_459 | ···················· | ···················· | ···················· | ···················· | ···················· | ···················· |
| C | 8905 hCoV-19/Morocco/6887/2020\|EPI_ISL_459965\|2020-03-0 | ···················· | ···················· | ···················· | ···················· | ···················· | ···················· |
| C | 8906 hCoV-19/Morocco/6888/2020\|EPI_ISL_459966\|2020-03-1 | ···················· | ···················· | ···················· | ···················· | ···················· | ···················· |
| C | 8907 hCoV-19/Morocco/6889/2020\|EPI_ISL_459967\|2020-03-1 | ···················· | ···················· | ···················· | ···················· | ···················· | ···················· |
| C | 8908 hCoV-19/Morocco/6890/2020\|EPI_ISL_459968\|2020-03-1 | ···················· | ···················· | ···················· | ···················· | ···················· | ···················· |
| C | 8909 hCoV-19/Morocco/6894/2020\|EPI_ISL_459972\|2020-03-2 | ···················· | ···················· | ···················· | ···················· | ···················· | ···················· |
| C | 8910 hCoV-19/Morocco/6895/2020\|EPI_ISL_459973\|2020-03-2 | ···················· | ···················· | ···················· | ···················· | ···················· | ···················· |
| C | 8911 hCoV-19/Morocco/6896/2020\|EPI_ISL_459974\|2020-03-2 | ···················· | ···················· | ···················· | ···················· | ···················· | ···················· |
| C | 8912 hCoV-19/Morocco/6897/2020\|EPI_ISL_459975\|2020-03-2 | ···················· | ···················· | ···················· | ···················· | ···················· | ···················· |
| C | 8913 hCoV-19/Morocco/6898/2020\|EPI_ISL_459976\|2020-03-1 | ···················· | ···················· | ···················· | ···················· | ···················· | ···················· |
| C | 8914 hCoV-19/Morocco/6899/2020\|EPI_ISL_459977\|2020-04-2 | ···················· | ···················· | ···················· | ···················· | ···················· | ···················· |
| C | 8915 hCoV-19/Morocco/6900/2020\|EPI_ISL_459978\|2020-04-2 | ···················· | ···················· | ···················· | ···················· | ···················· | ···················· |
| C | 8916 hCoV-19/Morocco/6901/2020\|EPI_ISL_459979\|2020-04-1 | ···················· | ···················· | ···················· | ···················· | ···················· | ···················· |
| C | 8917 hCoV-19/Morocco/6902/2020\|EPI_ISL_459980\|2020-04-1 | ···················· | ···················· | ···················· | ···················· | ···················· | ···················· |
| C | 8918 hCoV-19/Morocco/6903/2020\|EPI_ISL_459981\|2020-04-1 | ···················· | ···················· | ···················· | ···················· | ···················· | ···················· |
| C | 8919 hCoV-19/Morocco/6904/2020\|EPI_ISL_459982\|2020-04-1 | ···················· | ···················· | ···················· | ···················· | ···················· | ···················· |
| C | 8920 hCoV-19/Morocco/6905/2020\|EPI_ISL_459983\|2020-04-2 | ···················· | ···················· | ···················· | ···················· | ···················· | ···················· |
| C | 8921 hCoV-19/Morocco/6906/2020\|EPI_ISL_459984\|2020-04-0 | ···················· | ···················· | ···················· | ···················· | ···················· | ···················· |
| C | 8922 hCoV-19/Netherlands/Gelderland_40/2020\|EPI_ISL_460 | ···················· | ···················· | ···················· | ···················· | ···················· | ···················· |
| C | 8923 hCoV-19/Netherlands/Gelderland_41/2020\|EPI_ISL_460 | ···················· | ···················· | ···················· | ···················· | ···················· | ···················· |
| C | 8924 hCoV-19/Netherlands/Gelderland_42/2020\|EPI_ISL_460 | ···················· | ···················· | ···················· | ···················· | ···················· | ···················· |
| C | 8925 hCoV-19/Netherlands/Gelderland_50/2020\|EPI_ISL_460 | ···················· | ···················· | ···················· | ···················· | ···················· | ···················· |
| C | 8926 hCoV-19/Netherlands/Gelderland_52/2020\|EPI_ISL_460 | ···················· | ···················· | ···················· | ···················· | ···················· | ···················· |
| C | 8927 hCoV-19/Netherlands/Gelderland_53/2020\|EPI_ISL_460 | ···················· | ···················· | ···················· | ···················· | ···················· | ···················· |
| C | 8928 hCoV-19/Netherlands/Gelderland_54/2020\|EPI_ISL_460 | ···················· | ···················· | ···················· | ···················· | ···················· | ···················· |
| C | 8929 hCoV-19/Netherlands/Gelderland_56/2020\|EPI_ISL_460 | ···················· | ···················· | ···················· | ···················· | ···················· | ···················· |
| C | 8930 hCoV-19/Netherlands/Gelderland_57/2020\|EPI_ISL_460 | ···················· | ···················· | ···················· | ···················· | ···················· | ···················· |
| C | 8931 hCoV-19/Netherlands/Gelderland_60/2020\|EPI_ISL_460 | ···················· | ···················· | ···················· | ···················· | ···················· | ···················· |
| C | 8932 hCoV-19/Netherlands/Gelderland_61/2020\|EPI_ISL_460 | ···················· | ···················· | ···················· | ···················· | ···················· | ···················· |
| C | 8933 hCoV-19/Netherlands/Gelderland_62/2020\|EPI_ISL_460 | ···················· | ···················· | ···················· | ···················· | ···················· | ···················· |
| C | 8934 hCoV-19/Netherlands/Gelderland_63/2020\|EPI_ISL_460 | ···················· | ···················· | ···················· | ···················· | ···················· | ···················· |
| C | 8935 hCoV-19/Netherlands/Gelderland_64/2020\|EPI_ISL_460 | ···················· | ···················· | ···················· | ···················· | ···················· | ···················· |
| C | 8936 hCoV-19/Netherlands/Gelderland_65/2020\|EPI_ISL_460 | ···················· | ···················· | ···················· | ···················· | ···················· | ···················· |
| C | 8937 hCoV-19/Netherlands/Gelderland_66/2020\|EPI_ISL_460 | ···················· | ···················· | ···················· | ···················· | ···················· | ···················· |
| C | 8938 hCoV-19/Netherlands/Gelderland_67/2020\|EPI_ISL_460 | ···················· | ···················· | ···················· | ···················· | ···················· | ···················· |
| C | 8939 hCoV-19/Netherlands/Gelderland_68/2020\|EPI_ISL_460 | ···················· | ···················· | ···················· | ···················· | ···················· | ···················· |
| C | 8940 hCoV-19/Netherlands/Gelderland_69/2020\|EPI_ISL_460 | ···················· | ···················· | ···················· | ···················· | ···················· | ···················· |
| C | 8941 hCoV-19/Netherlands/Gelderland_70/2020\|EPI_ISL_460 | ···················· | ···················· | ···················· | ···················· | ···················· | ···················· |
| C | 8942 hCoV-19/Egypt/CUNCI-HGC008/2020\|EPI_ISL_468044\|202 | ···················· | ······A············· | ···················· | ···················· | ···················· | ···················· |
| C | 8943 hCoV-19/Egypt/CUNCI-HGC002/2020\|EPI_ISL_468045\|202 | ···················· | ······R············· | ···················· | ···················· | ···················· | ···················· |
| C | 8944 hCoV-19/Egypt/CUNCI-HGC007/2020\|EPI_ISL_468046\|202 | ···················· | ······A············· | ···················· | ···················· | ···················· | ···················· |
| C | 8945 hCoV-19/Egypt/CUNCI-HGC013/2020\|EPI_ISL_468047\|202 | ···················· | ···················· | ···················· | ···················· | ···················· | ···················· |
| C | 8946 hCoV-19/Egypt/CUNCI-HGC004/2020\|EPI_ISL_468048\|202 | ···················· | ···················· | ···················· | ···················· | ···················· | ···················· |
| C | 8947 hCoV-19/Egypt/CUNCI-HGC003/2020\|EPI_ISL_468049\|202 | ···················· | ······A············· | ···················· | ···················· | ···················· | ···················· |
| C | 8948 hCoV-19/Egypt/CUNCI-HGC006/2020\|EPI_ISL_468050\|202 | ···················· | ······A············· | ···················· | ···················· | ···················· | ···················· |
| C | 8949 hCoV-19/Egypt/CUNCI-HGC012/2020\|EPI_ISL_468051\|202 | ···················· | ···················· | ···················· | ···················· | ···················· | ···················· |
| C | 8950 hCoV-19/Egypt/CUNCI-HGC005/2020\|EPI_ISL_468052\|202 | ···················· | ···················· | ···················· | ···················· | ···················· | ···················· |
| C | 8951 hCoV-19/Egypt/CUNCI-HGC009/2020\|EPI_ISL_468053\|202 | ···················· | ···················· | ···················· | ···················· | ···················· | ···················· |
| C | 8952 hCoV-19/Egypt/CUNCI-HGC007-2/2020\|EPI_ISL_468054\|2 | ···················· | ······A············· | ···················· | ···················· | ···················· | ···················· |

| | | | | | | | | | |
|---|---|---|---|---|---|---|---|---|---|
| C | 8953 | hCoV-19/Egypt/CUNCI-HGC010/2020\|EPI_ISL_468055\|202 | ···················· | ···················· | ···················· | ···················· | ···················· | ···················· | |
| C | 8954 | hCoV-19/Egypt/CUNCI-HGC015/2020\|EPI_ISL_468056\|202 | ···················· | ···················· | ···················· | ···················· | ···················· | ···················· | |
| C | 8955 | hCoV-19/Egypt/CUNCI-HGC023/2020\|EPI_ISL_468057\|202 | ···················· | ······R············· | ···················· | ···················· | ···················· | ···················· | |
| C | 8956 | hCoV-19/Egypt/CUNCI-HGC016/2020\|EPI_ISL_468058\|202 | ···················· | ···················· | ···················· | ···················· | ···················· | ···················· | |
| C | 8957 | hCoV-19/Egypt/CUNCI-HGC011/2020\|EPI_ISL_468059\|202 | ···················· | ···················· | ···················· | ···················· | ···················· | ···················· | |
| C | 8958 | hCoV-19/Egypt/CUNCI-HGC014/2020\|EPI_ISL_468060\|202 | ···················· | ···················· | ···················· | ···················· | ···················· | ···················· | |
| C | 8959 | hCoV-19/Egypt/CUNCI-HGC021/2020\|EPI_ISL_468061\|202 | ···················· | ···················· | ···················· | ···················· | ···················· | ···················· | |
| C | 8960 | hCoV-19/Egypt/CUNCI-HGC028/2020\|EPI_ISL_468062\|202 | ···················· | ···················· | ···················· | ···················· | ···················· | ···················· | |
| C | 8961 | hCoV-19/England/CAMB-1AD0E6/2020\|EPI_ISL_447966\|20 | ···················· | ···················· | ···················· | ···················· | ···················· | ···················· | |
| C | 8962 | hCoV-19/England/CAMB-1AD101/2020\|EPI_ISL_447967\|20 | ···················· | ···················· | ···················· | ···················· | ···················· | ···················· | |
| C | 8963 | hCoV-19/England/CAMB-1AD13E/2020\|EPI_ISL_447969\|20 | ···················· | ···················· | ···················· | ···················· | ···················· | ···················· | |
| C | 8964 | hCoV-19/England/CAMB-1AD1E3/2020\|EPI_ISL_447977\|20 | ···················· | ···················· | ···················· | ···················· | ···················· | ···················· | |
| C | 8965 | hCoV-19/England/CAMB-1AD1F2/2020\|EPI_ISL_447978\|20 | ···················· | ···················· | ···················· | ···················· | ···················· | ···················· | |
| C | 8966 | hCoV-19/England/CAMB-1AD20E/2020\|EPI_ISL_447979\|20 | ···················· | ···················· | ···················· | ···················· | ···················· | ···················· | |
| C | 8967 | hCoV-19/England/CAMB-1AD23B/2020\|EPI_ISL_447980\|20 | ···················· | ···················· | ···················· | ···················· | ···················· | ···················· | |
| C | 8968 | hCoV-19/England/CAMB-1AD24A/2020\|EPI_ISL_447981\|20 | ···················· | ···················· | ···················· | ···················· | ···················· | ···················· | |
| C | 8969 | hCoV-19/England/CAMB-1AD268/2020\|EPI_ISL_447983\|20 | ···················· | ···················· | ···················· | ···················· | ···················· | ···················· | |
| C | 8970 | hCoV-19/England/CAMB-1AD277/2020\|EPI_ISL_447984\|20 | ···················· | ···················· | ···················· | ···················· | ···················· | ···················· | |
| C | 8971 | hCoV-19/England/CAMB-1AD2A4/2020\|EPI_ISL_447985\|20 | ···················· | ···················· | ···················· | ···················· | ···················· | ···················· | |
| C | 8972 | hCoV-19/England/CAMB-1AD2B3/2020\|EPI_ISL_447986\|20 | ···················· | ···················· | ···················· | ···················· | ···················· | ···················· | |
| C | 8973 | hCoV-19/England/CAMB-1AD2C2/2020\|EPI_ISL_447987\|20 | ···················· | ···················· | ···················· | ···················· | ···················· | ···················· | |
| C | 8974 | hCoV-19/England/NORW-E7791/2020\|EPI_ISL_449070\|202 | ···················· | ···················· | ···················· | ···················· | ···················· | ···················· | |
| C | 8975 | hCoV-19/England/NORW-E77CE/2020\|EPI_ISL_449071\|202 | ···················· | ···················· | ···················· | ···················· | ···················· | ···················· | |
| C | 8976 | hCoV-19/England/NORW-E77EC/2020\|EPI_ISL_449072\|202 | ···················· | ···················· | ···················· | ···················· | ···················· | ···················· | |
| C | 8977 | hCoV-19/England/NORW-E7816/2020\|EPI_ISL_449073\|202 | ···················· | ···················· | ···················· | ···················· | ···················· | ···················· | |
| C | 8978 | hCoV-19/England/NORW-E7825/2020\|EPI_ISL_449074\|202 | ···················· | ···················· | ···················· | ···················· | NNN················· | ···················· | 1 |
| C | 8979 | hCoV-19/England/NORW-E7834/2020\|EPI_ISL_449075\|202 | ···················· | ···················· | ···················· | ···················· | ···················· | ···················· | |
| C | 8980 | hCoV-19/England/NORW-E7861/2020\|EPI_ISL_449077\|202 | ···················· | ···················· | ···················· | ···················· | ···················· | ···················· | |
| C | 8981 | hCoV-19/England/NORW-E7870/2020\|EPI_ISL_449078\|202 | ···················· | ···················· | ···················· | ···················· | ···················· | ···················· | |
| C | 8982 | hCoV-19/England/NORW-E788F/2020\|EPI_ISL_449079\|202 | ···················· | ···················· | ···················· | ···················· | ···················· | ···················· | |
| C | 8983 | hCoV-19/England/NORW-E7DEA/2020\|EPI_ISL_449090\|202 | ···················· | ···················· | ···················· | ···················· | ···················· | ···················· | |
| C | 8984 | hCoV-19/England/NORW-E7DF9/2020\|EPI_ISL_449091\|202 | ···················· | ···················· | ···················· | ···················· | ···················· | ···················· | |
| C | 8985 | hCoV-19/England/NORW-E7E05/2020\|EPI_ISL_449092\|202 | ···················· | ···················· | ···················· | ···················· | ···················· | ···················· | |
| C | 8986 | hCoV-19/England/NORW-E7E9C/2020\|EPI_ISL_449093\|202 | ···················· | ···················· | ···················· | ···················· | ···················· | ···················· | |
| C | 8987 | hCoV-19/England/NORW-E7EAB/2020\|EPI_ISL_449094\|202 | ···················· | ···················· | ···················· | ···················· | ···················· | ···················· | |
| C | 8988 | hCoV-19/England/NORW-E7EBA/2020\|EPI_ISL_449095\|202 | ···················· | ···················· | ···················· | ···················· | ···················· | ···················· | |
| C | 8989 | hCoV-19/England/NORW-E7F4E/2020\|EPI_ISL_449096\|202 | ···················· | ···················· | ···················· | ···················· | ···················· | ···················· | |
| C | 8990 | hCoV-19/England/NORW-E7F5D/2020\|EPI_ISL_449097\|202 | ···················· | ···················· | ···················· | ···················· | ···················· | ···················· | |
| C | 8991 | hCoV-19/England/NORW-E8059/2020\|EPI_ISL_449099\|202 | ···················· | ···················· | ···················· | ···················· | ···················· | ···················· | |
| C | 8992 | hCoV-19/USA/NY-PV09384/2020\|EPI_ISL_450067\|2020-03 | ···················· | ···················· | ···················· | ···················· | ···················· | ···················· | |
| C | 8993 | hCoV-19/USA/NY-PV09385/2020\|EPI_ISL_450068\|2020-03 | ···················· | ···················· | ···················· | ···················· | ···················· | ···················· | |
| C | 8994 | hCoV-19/USA/NY-PV09387/2020\|EPI_ISL_450069\|2020-03 | ···················· | ···················· | ···················· | ···················· | ···················· | ···················· | |
| C | 8995 | hCoV-19/USA/NY-PV09388/2020\|EPI_ISL_450070\|2020-03 | ···················· | ···················A | ···················· | ···················· | ···················· | ···················· | 1 |
| C | 8996 | hCoV-19/USA/NY-PV09391/2020\|EPI_ISL_450072\|2020-03 | ···················· | ···················· | ···················· | ···················· | ···················· | ···················· | |
| C | 8997 | hCoV-19/USA/NY-PV09393/2020\|EPI_ISL_450074\|2020-03 | ···················· | ···················· | ···················· | ···················· | ···················· | ···················· | |
| C | 8998 | hCoV-19/USA/NY-PV09394/2020\|EPI_ISL_450075\|2020-03 | ···················· | ···················· | ···················· | ···················· | ···················· | ···················· | |
| C | 8999 | hCoV-19/USA/NY-PV09395/2020\|EPI_ISL_450076\|2020-03 | ···················· | ···················· | ···················· | ···················· | ···················· | ···················· | |
| C | 9000 | hCoV-19/USA/NY-PV09396/2020\|EPI_ISL_450077\|2020-03 | ···················· | ···················· | ···················· | ···················· | ···················· | ···················· | |
| C | 9001 | hCoV-19/USA/NY-PV09397/2020\|EPI_ISL_450078\|2020-03 | ···················· | ···················· | ···················· | ···················· | ···················· | ···················· | |
| C | 9002 | hCoV-19/USA/NY-PV09398/2020\|EPI_ISL_450079\|2020-03 | ···················· | ···················· | ···················· | ···················· | ···················· | ···················· | |
| C | 9003 | hCoV-19/USA/NY-PV09399/2020\|EPI_ISL_450080\|2020-03 | ···················· | ···················· | ···················· | ···················· | ···················· | ···················· | |
| C | 9004 | hCoV-19/USA/NY-PV09400/2020\|EPI_ISL_450081\|2020-03 | ···················· | ···················· | ···················· | ···················· | ···················· | ···················· | |
| C | 9005 | hCoV-19/USA/NY-PV09401/2020\|EPI_ISL_450082\|2020-03 | ···················· | ···················· | ···················· | ···················· | ···················· | ···················· | |
| C | 9006 | hCoV-19/USA/NY-PV09406/2020\|EPI_ISL_450086\|2020-03 | ···················· | ···················· | ···················· | ···················· | ···················· | ···················· | |
| C | 9007 | hCoV-19/USA/NY-PV09407/2020\|EPI_ISL_450087\|2020-03 | ···················· | ···················· | ···················· | ···················· | ···················· | ···················· | |
| C | 9008 | hCoV-19/USA/NY-PV09409/2020\|EPI_ISL_450089\|2020-03 | ···················· | ···················· | ···················· | ···················· | ···················· | ···················· | |
| C | 9009 | hCoV-19/USA/NY-PV09410/2020\|EPI_ISL_450090\|2020-03 | ···················· | ···················· | ···················· | ···················· | ···················· | ···················· | |
| C | 9010 | hCoV-19/USA/NY-PV09411/2020\|EPI_ISL_450091\|2020-03 | ···················· | ···················· | ···················· | ···················· | ···················· | ···················· | |
| C | 9011 | hCoV-19/USA/NY-PV09412/2020\|EPI_ISL_450092\|2020-03 | ···················· | ···················· | ···················· | ···················· | ···················· | ···················· | |
| C | 9012 | hCoV-19/USA/NY-PV09413/2020\|EPI_ISL_450093\|2020-03 | ···················· | ···················· | ···················· | ···················· | ···················· | ···················· | |
| C | 9013 | hCoV-19/USA/NY-PV09414/2020\|EPI_ISL_450094\|2020-03 | ···················· | ···················· | ···················· | ···················· | ···················· | ···················· | |
| C | 9014 | hCoV-19/USA/NY-PV09415/2020\|EPI_ISL_450095\|2020-03 | ···················· | ···················· | ···················· | ···················· | ···················· | ···················· | |
| C | 9015 | hCoV-19/USA/NY-PV09416/2020\|EPI_ISL_450096\|2020-03 | ···················· | ···················· | ···················· | ···················· | ···················· | ···················· | |
| C | 9016 | hCoV-19/USA/NY-PV09418/2020\|EPI_ISL_450097\|2020-03 | ···················· | ···················· | ···················· | ···················· | ···················· | ···················· | |
| C | 9017 | hCoV-19/USA/NY-PV09419/2020\|EPI_ISL_450098\|2020-03 | ···················· | ···················· | ···················· | ···················· | ···················· | ···················· | |
| C | 9018 | hCoV-19/USA/NY-PV09420/2020\|EPI_ISL_450099\|2020-03 | ···················· | ···················· | ···················· | ···················· | ···················· | ···················· | |
| C | 9019 | hCoV-19/Sichuan/SC-YB-085/2020\|EPI_ISL_451398\|2020 | ···················· | ···················· | ···················· | ···················· | ···················· | ···················· | |
| C | 9020 | hCoV-19/USA/LA-EVTL157/2020\|EPI_ISL_456000\|2020-04 | ···················· | ···················· | ···················· | ···················· | ···················· | ···················· | |
| C | 9021 | hCoV-19/USA/LA-EVTL158/2020\|EPI_ISL_456001\|2020-04 | ···················· | ···················· | ···················· | ···················· | ···················· | ···················· | |
| C | 9022 | hCoV-19/USA/LA-EVTL159/2020\|EPI_ISL_456002\|2020-04 | ···················· | ···················· | ···················· | ···················· | ···················· | ···················· | |
| C | 9023 | hCoV-19/USA/NY-NYUMC767/2020\|EPI_ISL_456004\|2020-0 | ···················· | ···················· | ···················· | ···················· | ···················· | ···················· | |
| C | 9024 | hCoV-19/USA/NY-NYUMC769/2020\|EPI_ISL_456006\|2020-0 | ···················· | ···················· | ···················· | ···················· | ···················· | ···················· | |
| C | 9025 | hCoV-19/USA/NY-NYUMC770/2020\|EPI_ISL_456007\|2020-0 | ···················· | ···················· | ···················· | ···················· | ···················· | ···················· | |
| C | 9026 | hCoV-19/USA/NY-NYUMC771/2020\|EPI_ISL_456008\|2020-0 | ···················· | ···················· | ···················· | ···················· | ···················· | ···················· | |
| C | 9027 | hCoV-19/USA/NY-NYUMC773/2020\|EPI_ISL_456010\|2020-0 | ···················· | ···················· | ···················· | ···················· | ···················· | ···················· | |
| C | 9028 | hCoV-19/USA/NY-NYUMC774/2020\|EPI_ISL_456011\|2020-0 | ···················· | ···················· | ···················· | ···················· | ···················· | ···················· | |
| C | 9029 | hCoV-19/USA/NY-NYUMC775/2020\|EPI_ISL_456012\|2020-0 | ···················· | ···················· | ···················· | ···················· | ···················· | ···················· | |
| C | 9030 | hCoV-19/USA/NY-NYUMC776/2020\|EPI_ISL_456013\|2020-0 | ···················· | ···················· | ···················· | ···················· | ···················· | ···················· | |

| | | | | | | | | |
|---|---|---|---|---|---|---|---|---|
| C | 9031 hCoV-19/USA/NY-NYUMC777/2020\|EPI_ISL_456014\|2020-0 | ···················· | ···················· | ···················· | ···················· | ···················· | ···················· | |
| C | 9032 hCoV-19/USA/NY-NYUMC778/2020\|EPI_ISL_456015\|2020-0 | ···················· | ···················· | ···················· | ···················· | ···················· | ···················· | |
| C | 9033 hCoV-19/USA/NY-NYUMC779/2020\|EPI_ISL_456016\|2020-0 | ···················· | ···················· | ···················· | ···················· | ···················· | ···················· | |
| C | 9034 hCoV-19/USA/NY-NYUMC780/2020\|EPI_ISL_456017\|2020-0 | ···················· | ···················· | ···················· | ···················· | ···················· | ···················· | |
| C | 9035 hCoV-19/USA/NY-NYUMC781/2020\|EPI_ISL_456018\|2020-0 | ···················· | ···················· | ···················· | ···················· | ···················· | ···················· | |
| C | 9036 hCoV-19/USA/NY-NYUMC782/2020\|EPI_ISL_456019\|2020-0 | ···················· | ···················· | ···················· | ···················· | ···················· | ···················· | |
| C | 9037 hCoV-19/USA/NY-NYUMC783/2020\|EPI_ISL_456020\|2020-0 | ···················· | ···················· | ···················· | ···················· | ···················· | ···················· | |
| C | 9038 hCoV-19/USA/NY-NYUMC784/2020\|EPI_ISL_456021\|2020-0 | ···················· | ···················· | ···················· | ···················· | ···················· | ···················· | |
| C | 9039 hCoV-19/USA/NY-NYUMC785/2020\|EPI_ISL_456022\|2020-0 | ···················· | ···················· | ···················· | ···················· | ···················· | ···················· | |
| C | 9040 hCoV-19/USA/NY-NYUMC786/2020\|EPI_ISL_456023\|2020-0 | ···················· | ···················· | ···················· | ···················· | ···················· | ···················· | |
| C | 9041 hCoV-19/USA/NY-NYUMC787/2020\|EPI_ISL_456024\|2020-0 | ···················· | ···················· | ···················· | ···················· | ···················· | ···················· | |
| C | 9042 hCoV-19/USA/NY-NYUMC788/2020\|EPI_ISL_456025\|2020-0 | ···················· | ···················· | ···················· | ···················· | ···················· | ···················· | |
| C | 9043 hCoV-19/USA/NY-NYUMC789/2020\|EPI_ISL_456026\|2020-0 | ···················· | ···················· | ···················· | ···················· | ···················· | ···················· | |
| C | 9044 hCoV-19/USA/NY-NYUMC790/2020\|EPI_ISL_456027\|2020-0 | ···················· | ···················· | ···················· | ···················· | ···················· | ···················· | |
| C | 9045 hCoV-19/USA/NY-NYUMC792/2020\|EPI_ISL_456028\|2020-0 | ···················· | ···················· | ···················· | ···················· | ···················· | ···················· | |
| C | 9046 hCoV-19/USA/NY-NYUMC793/2020\|EPI_ISL_456029\|2020-0 | ···················· | ···················· | ···················· | ···················· | ···················· | ···················· | |
| C | 9047 hCoV-19/USA/NY-NYUMC794/2020\|EPI_ISL_456030\|2020-0 | ···················· | ···················· | ···················· | ···················· | ···················· | ···················· | |
| C | 9048 hCoV-19/USA/NY-NYUMC795/2020\|EPI_ISL_456031\|2020-0 | ···················· | ···················· | ···················· | ···················· | ···················· | ···················· | |
| C | 9049 hCoV-19/USA/NY-NYUMC796/2020\|EPI_ISL_456032\|2020-0 | ···················· | ···················· | ···················· | ···················· | ···················· | ···················· | |
| C | 9050 hCoV-19/USA/NY-NYUMC797/2020\|EPI_ISL_456033\|2020-0 | ···················· | ···················· | ···················· | ···················· | ···················· | ···················· | |
| C | 9051 hCoV-19/USA/NY-NYUMC798/2020\|EPI_ISL_456034\|2020-0 | ···················· | ···················· | ···················· | ···················· | ···················· | ···················· | |
| C | 9052 hCoV-19/USA/NY-NYUMC799/2020\|EPI_ISL_456035\|2020-0 | ···················· | ···················· | ···················· | ···················· | ···················· | ···················· | |
| C | 9053 hCoV-19/USA/NY-NYUMC800/2020\|EPI_ISL_456036\|2020-0 | ···················· | ···················· | ···················· | ···················· | ···················· | ···················· | |
| C | 9054 hCoV-19/USA/NY-NYUMC801/2020\|EPI_ISL_456037\|2020-0 | ···················· | ···················· | ···················· | ···················· | ···················· | ···················· | |
| C | 9055 hCoV-19/USA/NY-NYUMC802/2020\|EPI_ISL_456038\|2020-0 | ···················· | ···················· | ···················· | ···················· | ···················· | ···················· | |
| C | 9056 hCoV-19/USA/NY-NYUMC803/2020\|EPI_ISL_456039\|2020-0 | ···················· | ···················· | ···················· | ···················· | ···················· | ···················· | |
| C | 9057 hCoV-19/England/LOND-D5B21/2020\|EPI_ISL_457301\|202 | ···················· | ···················· | ···················· | ···················· | ···················· | ···················· | |
| C | 9058 hCoV-19/England/NORW-EA136/2020\|EPI_ISL_457328\|202 | ···················· | ···················· | ···················· | ···················· | ···················· | ···················· | |
| C | 9059 hCoV-19/England/NORW-EA154/2020\|EPI_ISL_457329\|202 | ···················· | ···················· | ···················· | ···················· | ···················· | ···················· | |
| C | 9060 hCoV-19/England/NORW-EA172/2020\|EPI_ISL_457330\|202 | ···················· | ···················· | ···················· | ···················· | ···················· | ···················· | |
| C | 9061 hCoV-19/England/NORW-EA190/2020\|EPI_ISL_457331\|202 | ···················· | ···················· | ···················· | ···················· | ···················· | ···················· | |
| C | 9062 hCoV-19/England/NORW-EA1AF/2020\|EPI_ISL_457332\|202 | ···················· | ···················· | ···················· | ···················· | ···················· | ···················· | |
| C | 9063 hCoV-19/England/NORW-EA1BE/2020\|EPI_ISL_457333\|202 | ···················· | ···················· | ···················· | ···················· | ···················· | ···················· | |
| C | 9064 hCoV-19/England/NORW-EA224/2020\|EPI_ISL_457334\|202 | ···················· | ···················· | ···················· | ···················· | ···················· | ···················· | |
| C | 9065 hCoV-19/England/NORW-EA251/2020\|EPI_ISL_457335\|202 | ···················· | ···················· | ···················· | ···················· | ···················· | ···················· | |
| C | 9069 hCoV-19/England/NORW-EA2AC/2020\|EPI_ISL_457339\|202 | ···················· | ···················A | ···················· | ···················· | ···················· | ···················· | 1 |
| C | 9070 hCoV-19/England/NORW-EA303/2020\|EPI_ISL_457340\|202 | ···················· | ···················A | ···················· | ···················· | ···················· | ···················· | 1 |
| C | 9071 hCoV-19/England/NORW-EA38B/2020\|EPI_ISL_457341\|202 | ···················· | ···················A | ···················· | ···················· | ···················· | ···················· | 1 |
| C | 9072 hCoV-19/England/NORW-EA3C7/2020\|EPI_ISL_457342\|202 | ···················· | ···················· | ···················· | ···················· | ···················· | ···················· | |
| C | 9073 hCoV-19/England/NORW-EA400/2020\|EPI_ISL_457344\|202 | ···················· | ···················· | ···················· | ···················· | ···················· | ···················· | |
| C | 9074 hCoV-19/England/NORW-EA41F/2020\|EPI_ISL_457345\|202 | ···················· | ···················A | ···················· | ···················· | ···················· | ···················· | 1 |
| C | 9075 hCoV-19/England/NORW-EA42E/2020\|EPI_ISL_457346\|202 | ···················· | ···················· | ···················· | ···················· | ···················· | ···················· | |
| C | 9076 hCoV-19/England/NORW-EA46A/2020\|EPI_ISL_457347\|202 | ···················· | ···················A | ···················· | ···················· | ···················· | ···················· | 1 |
| C | 9079 hCoV-19/England/NORW-EAAA4/2020\|EPI_ISL_457352\|202 | ···················· | ···················· | ···················· | ···················· | ···················· | ···················· | |
| C | 9081 hCoV-19/England/NORW-EAB38/2020\|EPI_ISL_457354\|202 | ···················· | ···················· | ··········T········· | ···················· | ···················· | ···················· | |
| C | 9082 hCoV-19/England/NORW-EAB47/2020\|EPI_ISL_457355\|202 | ···················· | ···················· | ··········T········· | ···················· | ···················· | ···················· | |
| C | 9083 hCoV-19/England/NORW-EAB56/2020\|EPI_ISL_457356\|202 | ···················· | ···················· | ··········T········· | ···················· | ···················· | ···················· | |
| C | 9084 hCoV-19/England/NORW-EAB74/2020\|EPI_ISL_457357\|202 | ···················· | ···················· | ···················· | ···················· | ···················· | ···················· | |
| C | 9085 hCoV-19/England/NORW-EAB92/2020\|EPI_ISL_457358\|202 | ···················· | ···················· | ···················· | ···················· | ···················· | ···················· | |
| C | 9086 hCoV-19/England/NORW-EABDE/2020\|EPI_ISL_457360\|202 | ···················· | ···················· | ···················· | ···················· | ···················· | ···················· | |
| C | 9087 hCoV-19/England/NORW-EABED/2020\|EPI_ISL_457361\|202 | ···················· | ···················· | ···················· | ···················· | ···················· | ···················· | |
| C | 9088 hCoV-19/England/NORW-EAC08/2020\|EPI_ISL_457362\|202 | ···················· | ···················· | ···················· | ···················· | ···················· | ···················· | |
| C | 9089 hCoV-19/England/NORW-EAC35/2020\|EPI_ISL_457365\|202 | ···················· | ···················· | ···················· | ···················· | ···················· | ···················· | |
| C | 9091 hCoV-19/England/NORW-EAC80/2020\|EPI_ISL_457367\|202 | ···················· | ···················· | ···················· | ···················· | ···················· | ···················· | |
| C | 9092 hCoV-19/England/NORW-EACBD/2020\|EPI_ISL_457368\|202 | ··················T· | ···················· | ···················· | ···················· | ···················· | ···················· | 1 |
| C | 9093 hCoV-19/England/NORW-EACCC/2020\|EPI_ISL_457369\|202 | ···················· | ···················· | ···················· | ···················· | ···················· | ···················· | |
| C | 9094 hCoV-19/England/NORT-289155/2020\|EPI_ISL_458614\|20 | ···················· | ···················· | ···················· | ···················· | ···················· | ···················· | |
| C | 9095 hCoV-19/England/NORT-289313/2020\|EPI_ISL_458615\|20 | ···················· | ···················· | ···················· | ···················· | ···················· | ···················· | |
| C | 9096 hCoV-19/England/NORT-2897CC/2020\|EPI_ISL_458616\|20 | ···················· | ···················· | ···················· | ···················· | ···················· | ···················· | |
| C | 9097 hCoV-19/England/NORT-2891DD/2020\|EPI_ISL_458617\|20 | ···················· | ···················· | ···················· | ···················· | ···················· | ···················· | |
| C | 9098 hCoV-19/England/NORT-289234/2020\|EPI_ISL_458618\|20 | ···················· | ···················· | ···················· | ···················· | ···················· | ···················· | |
| C | 9099 hCoV-19/England/NORT-288F4D/2020\|EPI_ISL_458619\|20 | ···················· | ···················· | ···················· | ···················· | ···················· | ···················· | |
| C | 9100 hCoV-19/England/NORT-28960B/2020\|EPI_ISL_458620\|20 | ···················· | ···················· | ···················· | ···················· | ···················· | ···················· | |
| C | 9101 hCoV-19/England/NORT-289094/2020\|EPI_ISL_458621\|20 | ···················· | ···················· | ···················· | ···················· | ···················· | ···················· | |
| C | 9102 hCoV-19/England/NORT-288FD4/2020\|EPI_ISL_458622\|20 | ···················· | ···················· | ···················· | ···················· | ···················· | ···················· | |
| C | 9103 hCoV-19/England/NORT-289067/2020\|EPI_ISL_458623\|20 | ···················· | ···················· | ···················· | ···················· | ···················· | ···················· | |
| C | 9104 hCoV-19/England/NORT-289735/2020\|EPI_ISL_458624\|20 | ···················· | ···················· | ···················· | ···················· | ···················· | ···················· | |
| C | 9105 hCoV-19/England/NORT-2890EF/2020\|EPI_ISL_458660\|20 | ···················· | ···················· | ···················· | ···················· | ···················· | ···················· | |
| C | 9106 hCoV-19/England/NORT-289252/2020\|EPI_ISL_458661\|20 | ···················· | ···················· | ···················· | ···················· | ···················· | ···················· | |
| C | 9107 hCoV-19/England/NORT-2894D4/2020\|EPI_ISL_458662\|20 | ···················· | ···················· | ···················· | ···················· | ···················· | ···················· | |
| C | 9108 hCoV-19/England/NORT-289656/2020\|EPI_ISL_458664\|20 | ···················· | ···················· | ···················· | ···················· | ···················· | ···················· | |
| C | 9109 hCoV-19/England/NORT-28945C/2020\|EPI_ISL_458667\|20 | ···················· | ···················· | ···················· | ···················· | ···················· | ···················· | |
| C | 9110 hCoV-19/England/NORT-288FC5/2020\|EPI_ISL_458668\|20 | ···················· | ···················· | ···················· | ···················· | ···················· | ···················· | |
| C | 9111 hCoV-19/England/NORT-288DAD/2020\|EPI_ISL_458669\|20 | ···················· | ···················· | ···················· | ···················· | ···················· | ···················· | |
| C | 9112 hCoV-19/England/NORT-289270/2020\|EPI_ISL_458670\|20 | ···················· | ···················· | ···················· | ···················· | ···················· | ···················· | |
| C | 9113 hCoV-19/England/NORT-289780/2020\|EPI_ISL_458671\|20 | ···················· | ···················· | ···················· | ···················· | ···················· | ···················· | |
| C | 9114 hCoV-19/England/NORT-289999/2020\|EPI_ISL_458672\|20 | ···················· | ···················· | ···················· | ···················· | ···················· | ···················· | |
| C | 9115 hCoV-19/England/NORT-28936E/2020\|EPI_ISL_458674\|20 | ···················· | ···················· | ···················· | ···················· | ···················· | ···················· | |

| | | | | | | | | |
|---|---|---|---|---|---|---|---|---|
| C | 9116 hCoV-19/England/NORT-2898BA/2020\|EPI_ISL_458675\|20 | .................... | .................... | .................... | .................... | .................... | .................... | |
| C | 9117 hCoV-19/England/NORT-28993F/2020\|EPI_ISL_458676\|20 | .................... | .................... | .................... | .................... | .................... | .................... | |
| C | 9118 hCoV-19/England/NORT-2891BF/2020\|EPI_ISL_458677\|20 | .................... | .................... | .................... | .................... | .................... | .................... | |
| C | 9119 hCoV-19/England/NORT-289164/2020\|EPI_ISL_458678\|20 | .................... | .................... | .................... | .................... | .................... | .................... | |
| C | 9120 hCoV-19/England/NORT-289753/2020\|EPI_ISL_458679\|20 | .................... | .................... | .................... | .................... | .................... | .................... | |
| C | 9121 hCoV-19/England/NORT-289814/2020\|EPI_ISL_458681\|20 | .................... | ...................A | .................... | .................... | .................... | .................... | 1 |
| C | 9122 hCoV-19/England/NORT-288F6B/2020\|EPI_ISL_458684\|20 | .................... | .................... | .................... | .................... | .................... | .................... | |
| C | 9123 hCoV-19/England/NORT-28910A/2020\|EPI_ISL_458686\|20 | .................... | .................... | .................... | .................... | .................... | .................... | |
| C | 9124 hCoV-19/England/NORT-289173/2020\|EPI_ISL_458687\|20 | .................... | .................... | .................... | .................... | .................... | .................... | |
| C | 9125 hCoV-19/England/NORT-289119/2020\|EPI_ISL_458688\|20 | .................... | .................... | .................... | .................... | .................... | .................... | |
| C | 9126 hCoV-19/England/NORT-2896DE/2020\|EPI_ISL_458689\|20 | .................... | .................... | .................... | .................... | .................... | .................... | |
| C | 9127 hCoV-19/England/NORT-289058/2020\|EPI_ISL_458690\|20 | .................... | .................... | .................... | .................... | .................... | .................... | |
| C | 9128 hCoV-19/England/NORT-289489/2020\|EPI_ISL_458691\|20 | .................... | .................... | .................... | .................... | .................... | .................... | |
| C | 9129 hCoV-19/England/NORT-28937D/2020\|EPI_ISL_458693\|20 | .................... | .................... | .................... | .................... | .................... | .................... | |
| C | 9130 hCoV-19/England/NORT-289137/2020\|EPI_ISL_458694\|20 | .................... | .................... | .................... | .................... | .................... | .................... | |
| C | 9131 hCoV-19/England/NORT-28902B/2020\|EPI_ISL_458696\|20 | .................... | .................... | .................... | .................... | .................... | .................... | |
| C | 9132 hCoV-19/England/NORT-28946B/2020\|EPI_ISL_458697\|20 | .................... | .................... | .................... | .................... | .................... | .................... | |
| C | 9133 hCoV-19/England/NORT-2893AA/2020\|EPI_ISL_458699\|20 | .................... | .................... | .................... | .................... | .................... | .................... | |
| C | 9134 hCoV-19/England/BRIS-129826/2020\|EPI_ISL_458801\|20 | .................... | .................... | .................... | .................... | .................... | .................... | |
| C | 9135 hCoV-19/England/BRIS-12903D/2020\|EPI_ISL_458803\|20 | .................... | .................... | .................... | .................... | .................... | .................... | |
| C | 9136 hCoV-19/England/BRIS-128836/2020\|EPI_ISL_458804\|20 | .................... | .................... | .................... | .................... | .................... | .................... | |
| C | 9137 hCoV-19/England/BRIS-12960E/2020\|EPI_ISL_458808\|20 | .................... | .................... | .................... | .................... | .................... | .................... | |
| C | 9138 hCoV-19/England/BRIS-12962C/2020\|EPI_ISL_458810\|20 | .................... | .................... | .................... | .................... | .................... | .................... | |
| C | 9139 hCoV-19/England/BRIS-128F5F/2020\|EPI_ISL_458811\|20 | .................... | .................... | .................... | .................... | .................... | .................... | |
| C | 9140 hCoV-19/Malaysia/IMR_WC2665/2020\|EPI_ISL_459955\|20 | .................... | .................... | .................... | .................... | .................... | .................... | |
| C | 9141 hCoV-19/USA/MI-MDHHS-SC20455/2020\|EPI_ISL_459994\|2 | .................... | .................... | .................... | .................... | .................... | .................... | |
| C | 9142 hCoV-19/USA/MI-MDHHS-SC20456/2020\|EPI_ISL_459995\|2 | .................... | .................... | .................... | .................... | .................... | .................... | |
| C | 9143 hCoV-19/USA/MI-MDHHS-SC20457/2020\|EPI_ISL_459996\|2 | .................... | .................... | .................... | .................... | .................... | .................... | |
| C | 9144 hCoV-19/USA/MI-MDHHS-SC20458/2020\|EPI_ISL_459997\|2 | .................... | .................... | .................... | .................... | .................... | .................... | |
| C | 9145 hCoV-19/USA/MI-MDHHS-SC20459/2020\|EPI_ISL_459998\|2 | .................... | .................... | .................... | .................... | .................... | .................... | |
| C | 9146 hCoV-19/USA/MI-MDHHS-SC20460/2020\|EPI_ISL_459999\|2 | .................... | .................... | .................... | .................... | .................... | .................... | |
| C | 9147 hCoV-19/Netherlands/Gelderland_30/2020\|EPI_ISL_460 | .................... | .................... | .................... | .................... | .................... | .................... | |
| C | 9148 hCoV-19/Netherlands/Gelderland_31/2020\|EPI_ISL_460 | .................... | .................... | .................... | .................... | .................... | .................... | |
| C | 9149 hCoV-19/Netherlands/Gelderland_32/2020\|EPI_ISL_460 | .................... | .................... | .................... | .................... | .................... | .................... | |
| C | 9150 hCoV-19/Netherlands/Gelderland_33/2020\|EPI_ISL_460 | .................... | .................... | .................... | .................... | .................... | .................... | |
| C | 9151 hCoV-19/Netherlands/Gelderland_34/2020\|EPI_ISL_460 | .................... | .................... | .................... | .................... | .................... | .................... | |
| C | 9152 hCoV-19/Netherlands/Gelderland_35/2020\|EPI_ISL_460 | .................... | .................... | .................... | .................... | .................... | .................... | |
| C | 9153 hCoV-19/Netherlands/Gelderland_36/2020\|EPI_ISL_460 | .................... | .................... | .................... | .................... | .................... | .................... | |
| C | 9154 hCoV-19/Netherlands/Gelderland_37/2020\|EPI_ISL_460 | .................... | .................... | .................... | .................... | .................... | .................... | |
| C | 9155 hCoV-19/Netherlands/Gelderland_38/2020\|EPI_ISL_460 | .................... | .................... | .................... | .................... | .................... | .................... | |
| C | 9156 hCoV-19/Netherlands/Gelderland_39/2020\|EPI_ISL_460 | .................... | .................... | .................... | ..............A..... | .................... | .................... | |
| C | 9157 hCoV-19/Netherlands/Gelderland_43/2020\|EPI_ISL_460 | .................... | .................... | .................... | .................... | .................... | .................... | |
| C | 9158 hCoV-19/Netherlands/Gelderland_44/2020\|EPI_ISL_460 | .................... | .................... | .................... | .................... | .................... | .................... | |
| C | 9159 hCoV-19/Netherlands/Gelderland_45/2020\|EPI_ISL_460 | .................... | .................... | .................... | .................... | .................... | .................... | |
| C | 9160 hCoV-19/Netherlands/Gelderland_46/2020\|EPI_ISL_460 | .................... | .................... | .................... | .................... | .................... | .................... | |
| C | 9161 hCoV-19/Netherlands/Gelderland_47/2020\|EPI_ISL_460 | .................... | .................... | .................... | .................... | .................... | .................... | |
| C | 9162 hCoV-19/Netherlands/Gelderland_48/2020\|EPI_ISL_460 | .................... | .................... | .................... | .................... | .................... | .................... | |
| C | 9163 hCoV-19/Netherlands/Gelderland_49/2020\|EPI_ISL_460 | .................... | .................... | .................... | .................... | .................... | .................... | |
| C | 9164 hCoV-19/Netherlands/Gelderland_71/2020\|EPI_ISL_460 | .................... | .................... | .................... | .................... | .................... | .................... | |
| C | 9165 hCoV-19/Netherlands/Gelderland_72/2020\|EPI_ISL_460 | .................... | .................... | .................... | .................... | .................... | .................... | |
| C | 9166 hCoV-19/Netherlands/Gelderland_73/2020\|EPI_ISL_460 | .................... | .................... | .................... | .................... | .................... | .................... | |
| C | 9167 hCoV-19/Netherlands/Gelderland_74/2020\|EPI_ISL_460 | .................... | .................... | .................... | .................... | .................... | .................... | |
| C | 9168 hCoV-19/Netherlands/Gelderland_75/2020\|EPI_ISL_460 | .................... | .................... | .................... | .................... | .................... | .................... | |
| C | 9169 hCoV-19/Netherlands/Gelderland_76/2020\|EPI_ISL_460 | .................... | .................... | .................... | .................... | .................... | .................... | |
| C | 9170 hCoV-19/Netherlands/Gelderland_77/2020\|EPI_ISL_460 | .................... | .................... | .................... | .................... | .................... | .................... | |
| C | 9171 hCoV-19/Netherlands/Gelderland_78/2020\|EPI_ISL_460 | .................... | .................... | .................... | .................... | .................... | .................... | |
| C | 9172 hCoV-19/Netherlands/Gelderland_79/2020\|EPI_ISL_460 | .................... | .................... | .................... | .................... | .................... | .................... | |
| C | 9173 hCoV-19/Netherlands/Gelderland_80/2020\|EPI_ISL_460 | .................... | .................... | .................... | .................... | .................... | .................... | |
| C | 9174 hCoV-19/Netherlands/Gelderland_81/2020\|EPI_ISL_460 | .................... | .................... | .................... | .................... | .................... | .................... | |
| C | 9175 hCoV-19/Netherlands/Gelderland_82/2020\|EPI_ISL_460 | .................... | .................... | .................... | .................... | .................... | .................... | |
| C | 9176 hCoV-19/Netherlands/Gelderland_83/2020\|EPI_ISL_460 | .................... | .................... | .................... | .................... | .................... | .................... | |
| C | 9177 hCoV-19/Netherlands/Gelderland_84/2020\|EPI_ISL_460 | .................... | .................... | .................... | .................... | .................... | .................... | |
| C | 9178 hCoV-19/Netherlands/Gelderland_85/2020\|EPI_ISL_460 | .................... | .................... | .................... | .................... | .................... | .................... | |
| C | 9179 hCoV-19/Netherlands/Gelderland_86/2020\|EPI_ISL_460 | .................... | .................... | .................... | .................... | .................... | .................... | |
| C | 9180 hCoV-19/Netherlands/Gelderland_87/2020\|EPI_ISL_460 | .................... | .................... | .................... | .................... | .................... | .................... | |
| C | 9181 hCoV-19/Netherlands/Gelderland_88/2020\|EPI_ISL_460 | .................... | .................... | .................... | .................... | .................... | .................... | |
| C | 9182 hCoV-19/Netherlands/Gelderland_90/2020\|EPI_ISL_460 | .................... | .................... | .................... | .................... | .................... | .................... | |
| C | 9183 hCoV-19/Netherlands/Gelderland_91/2020\|EPI_ISL_460 | .................... | .................... | .................... | .................... | .................... | .................... | |
| C | 9184 hCoV-19/Netherlands/Gelderland_93/2020\|EPI_ISL_460 | .................... | .................... | .................... | .................... | .................... | .................... | |
| C | 9185 hCoV-19/Netherlands/Gelderland_94/2020\|EPI_ISL_460 | .................... | .................... | .................... | .................... | .................... | .................... | |
| C | 9186 hCoV-19/Netherlands/Gelderland_95/2020\|EPI_ISL_460 | .................... | .................... | .................... | .................... | .................... | .................... | |
| C | 9187 hCoV-19/Netherlands/Gelderland_96/2020\|EPI_ISL_460 | .................... | .................... | .................... | .................... | .................... | .................... | |
| C | 9188 hCoV-19/Netherlands/Gelderland_97/2020\|EPI_ISL_460 | .................... | .................... | .................... | .................... | .................... | .................... | |
| C | 9189 hCoV-19/Netherlands/Gelderland_98/2020\|EPI_ISL_460 | .................... | .................... | .................... | .................... | .................... | .................... | |
| C | 9190 hCoV-19/Netherlands/Gelderland_99/2020\|EPI_ISL_460 | .................... | .................... | .................... | .................... | .................... | .................... | |
| C | 9191 hCoV-19/Netherlands/Limburg_11/2020\|EPI_ISL_460999 | .................... | .................... | .................... | .................... | .................... | .................... | |
| C | 9192 hCoV-19/England/SHEF-CAA8C/2020\|EPI_ISL_462046\|202 | .................... | .................... | .................... | .................... | .................... | .................... | |
| C | 9193 hCoV-19/England/SHEF-CAF05/2020\|EPI_ISL_462047\|202 | .................... | .................... | .................... | .................... | .................... | .................... | |

C 9194 hCoV-19/England/SHEF-CB0E3/2020|EPI_ISL_462048|202 ···················· ···················· ····················· ···················· ···················· ····················
C 9195 hCoV-19/England/SHEF-CB49C/2020|EPI_ISL_462049|202 ···················· ···················· ····················· ···················· ···················· ····················
C 9196 hCoV-19/England/SHEF-CB739/2020|EPI_ISL_462050|202 ···················· ···················· ····················· ···················· ···················· ····················
C 9197 hCoV-19/England/SHEF-CB933/2020|EPI_ISL_462051|202 ···················· ···················· ····················· ···················· ···················· ····················
C 9198 hCoV-19/England/SHEF-CBB6A/2020|EPI_ISL_462052|202 ···················· ···················· ····················· ···················· ···················· ····················
C 9199 hCoV-19/England/SHEF-CBB97/2020|EPI_ISL_462053|202 ···················· ···················· ····················· ···················· ···················· ····················
C 9200 hCoV-19/England/SHEF-CBF22/2020|EPI_ISL_462054|202 ···················· ···················· ····················· ···················· ···················· ····················
C 9201 hCoV-19/England/SHEF-CC8DB/2020|EPI_ISL_462056|202 ···················· ···················· ····················· ···················· ···················· ····················
C 9202 hCoV-19/England/SHEF-CCAE4/2020|EPI_ISL_462058|202 ···················· ···················· ····················· ···················· ···················· ····················
C 9203 hCoV-19/England/SHEF-CCD54/2020|EPI_ISL_462059|202 ···················· ···················· ····················· ···················· ···················· ····················
C 9204 hCoV-19/England/SHEF-CCECA/2020|EPI_ISL_462060|202 ···················· ···················· ····················· ···················· ···················· ····················
C 9205 hCoV-19/England/SHEF-CCEE8/2020|EPI_ISL_462061|202 ···················· ···················· ····················· ···················· ···················· ····················
C 9206 hCoV-19/England/SHEF-CD96E/2020|EPI_ISL_462064|202 ···················· ···················· ····················· ···················· ···················· ····················
C 9207 hCoV-19/England/SHEF-CDD08/2020|EPI_ISL_462065|202 ···················· ···················· ····················· ···················· ···················· ····················
C 9208 hCoV-19/England/SHEF-CE04A/2020|EPI_ISL_462066|202 ···················· ···················· ····················· ···················· ···················· ····················
C 9209 hCoV-19/England/SHEF-CE62A/2020|EPI_ISL_462067|202 ···················· ···················· ····················· ···················· ···················· ····················
C 9210 hCoV-19/England/SHEF-CF39B/2020|EPI_ISL_462069|202 ···················· ···················· ····················· ···················· ···················· ····················
C 9211 hCoV-19/England/SHEF-CF595/2020|EPI_ISL_462070|202 ···················· ···················· ····················· ···················· ···················· ····················
C 9212 hCoV-19/England/SHEF-D04EF/2020|EPI_ISL_462072|202 ···················· ····A··············· ····················· ···················· ···················· ····················
C 9213 hCoV-19/England/SHEF-D086B/2020|EPI_ISL_462074|202 ···················· ···················· ····················· ···················· ···················· ····················
C 9214 hCoV-19/England/SHEF-D0959/2020|EPI_ISL_462075|202 ···················· ···················· ····················· ···················· ···················· ····················
C 9215 hCoV-19/England/SHEF-D0B71/2020|EPI_ISL_462076|202 ···················· ···················· ····················· ···················· ···················· ····················
C 9216 hCoV-19/England/SHEF-D1606/2020|EPI_ISL_462078|202 ···················· ···················· ····················· ···················· ···················· ····················
C 9217 hCoV-19/England/NORW-E7078/2020|EPI_ISL_449001|202 ···················· ···················· ····················· ···················· ···················· ····················
C 9218 hCoV-19/England/NORW-E70E1/2020|EPI_ISL_449002|202 ···················· ···················· ····················· ···················· ···················· ····················
C 9219 hCoV-19/England/NORW-E711B/2020|EPI_ISL_449003|202 ···················· ···················· ····················· ···················· ···················· ····················
C 9221 hCoV-19/England/NORW-E7139/2020|EPI_ISL_449005|202 ···················· ···················· ····················· ···················· ···················· ····················
C 9222 hCoV-19/England/NORW-E7148/2020|EPI_ISL_449006|202 ···················· ···················· ····················· ···················· ···················· ····················
C 9223 hCoV-19/England/NORW-E7166/2020|EPI_ISL_449007|202 ···················· ···················· ····················· ···················· ···················· ····················
C 9224 hCoV-19/England/NORW-E7175/2020|EPI_ISL_449008|202 ···················· ···················· ····················· ···················· ···················· ····················
C 9225 hCoV-19/England/NORW-E7193/2020|EPI_ISL_449009|202 ···················· ···················· ····················· ···················· ···················· ····················
C 9226 hCoV-19/England/NORW-E71A2/2020|EPI_ISL_449010|202 ···················· ···················· ····················· ···················· ···················· ····················
C 9227 hCoV-19/England/NORW-E71B1/2020|EPI_ISL_449011|202 ···················· ···················· ····················· ···················· ···················· ····················
C 9228 hCoV-19/England/NORW-E71C0/2020|EPI_ISL_449012|202 ···················· ···················· ····················· ···················· ···················· ····················
C 9229 hCoV-19/England/NORW-E7227/2020|EPI_ISL_449013|202 ···················· ···················· ····················· ···················· ···················· ····················
C 9230 hCoV-19/England/NORW-E7272/2020|EPI_ISL_449015|202 ···················· ···················· ····················· ···················· ···················· ····················
C 9232 hCoV-19/England/NORW-E72CD/2020|EPI_ISL_449019|202 ···················· ···················· ····················· ···················· ···················· ····················
C 9233 hCoV-19/England/NORW-E72DC/2020|EPI_ISL_449020|202 ···················· ···················· ····················· ···················· ···················· ····················
C 9234 hCoV-19/England/NORW-E72FA/2020|EPI_ISL_449021|202 ···················· ···················· ····················· ···················· ···················· ····················
C 9235 hCoV-19/England/NORW-E7306/2020|EPI_ISL_449022|202 ···················· ···················· ····················· ···················· ···················· ····················
C 9236 hCoV-19/England/NORW-E7315/2020|EPI_ISL_449023|202 ···················· ···················· ····················· ···················· ···················· ····················
C 9237 hCoV-19/England/NORW-E7324/2020|EPI_ISL_449024|202 ···················· ···················· ····················· ···················· ···················· ····················
C 9238 hCoV-19/England/NORW-E7333/2020|EPI_ISL_449025|202 ···················· ···················· ····················· ···················· ···················· ····················
C 9239 hCoV-19/England/NORW-E7351/2020|EPI_ISL_449026|202 ···················· ···················· ····················· ···················· ···················· ····················
C 9241 hCoV-19/England/NORW-E737F/2020|EPI_ISL_449028|202 ···················· ···················· ····················· ···················· ···················· ····················
C 9242 hCoV-19/England/NORW-E738E/2020|EPI_ISL_449029|202 ···················· ···················· ····················· ···················· ···················· ····················
C 9243 hCoV-19/England/NORW-E7588/2020|EPI_ISL_449050|202 ···················· ···················· ····················· ···················· ···················· ····················
C 9244 hCoV-19/England/NORW-E75A6/2020|EPI_ISL_449051|202 ···················· ···················· ····················· ···················· ···················· ····················
C 9245 hCoV-19/England/NORW-E75C4/2020|EPI_ISL_449053|202 ···················· ···················· ····················· ···················· ···················· ····················
C 9246 hCoV-19/England/NORW-E75D3/2020|EPI_ISL_449054|202 ···················· ···················· ····················· ···················· ···················· ····················
C 9247 hCoV-19/England/NORW-E75E2/2020|EPI_ISL_449055|202 ···················· ···················· ····················· ···················· ···················· ····················
C 9248 hCoV-19/England/NORW-E762B/2020|EPI_ISL_449056|202 ···················· ···················· ····················· ···················· ···················· ····················
C 9249 hCoV-19/England/NORW-E7649/2020|EPI_ISL_449057|202 ···················· ···················· ····················· ···················· ···················· ····················
C 9250 hCoV-19/England/NORW-E7667/2020|EPI_ISL_449059|202 ···················· ···················· ····················· ···················· ···················· ····················
C 9251 hCoV-19/England/NORW-E789E/2020|EPI_ISL_449080|202 ···················· ···················· ····················· ···················· ···················· ····················
C 9252 hCoV-19/England/NORW-E78BC/2020|EPI_ISL_449081|202 ···················· ···················· ····················· ···················· ···················· ····················
C 9253 hCoV-19/England/NORW-E7922/2020|EPI_ISL_449082|202 ···················· ···················· ····················· ···················· ···················· ····················
C 9254 hCoV-19/England/NORW-E7931/2020|EPI_ISL_449083|202 ···················· ···················· ····················· ···················· ···················· ····················
C 9255 hCoV-19/England/NORW-E7940/2020|EPI_ISL_449084|202 ···················· ···················· ····················· ···················· ···················· ····················
C 9256 hCoV-19/England/NORW-E797D/2020|EPI_ISL_449085|202 ···················· ···················· ····················· ···················· ···················· ····················
C 9257 hCoV-19/England/NORW-E79D7/2020|EPI_ISL_449086|202 ···················· ···················· ····················· ···················· ···················· ····················
C 9258 hCoV-19/England/NORW-E79E6/2020|EPI_ISL_449087|202 ···················· ···················· ····················· ···················· ···················· ····················
C 9260 hCoV-19/USA/WA-S830/2020|EPI_ISL_450000|2020-04-16 ···················· ···················· ····················· ···················· ···················· ····················
C 9261 hCoV-19/USA/WA-S831/2020|EPI_ISL_450001|2020-04-16 ···················· ···················· ····················· ···················· ···················· ····················
C 9262 hCoV-19/USA/WA-S832/2020|EPI_ISL_450002|2020-04-16 ···················· ···················· ····················· ···················· ···················· ····················
C 9263 hCoV-19/USA/WA-S833/2020|EPI_ISL_450003|2020-04-16 ···················· ···················· ····················· ···················· ···················· ····················
C 9264 hCoV-19/USA/NY-PV09312/2020|EPI_ISL_450004|2020-03 ···················· ···················· ····················· ···················· ···················· ····················
C 9265 hCoV-19/USA/NY-PV09313/2020|EPI_ISL_450005|2020-03 ···················· ···················· ····················· ···················· ···················· ····················
C 9266 hCoV-19/USA/NY-PV09314/2020|EPI_ISL_450006|2020-03 ···················· ···················· ····················· ···················· ···················· ····················
C 9267 hCoV-19/USA/NY-PV09315/2020|EPI_ISL_450007|2020-03 ···················· ···················· ····················· ···················· ···················· ····················
C 9268 hCoV-19/USA/NY-PV09316/2020|EPI_ISL_450008|2020-03 ···················· ···················· ····················· ···················· ···················· ····················
C 9269 hCoV-19/USA/NY-PV09317/2020|EPI_ISL_450009|2020-03 ···················· ···················· ····················· ···················· ···················· ····················
C 9270 hCoV-19/USA/NY-PV09318/2020|EPI_ISL_450010|2020-03 ···················· ···················· ····················· ···················· ···················· ····················
C 9271 hCoV-19/USA/NY-PV09322/2020|EPI_ISL_450011|2020-03 ···················· ···················· ····················· ···················· ···················· ····················
C 9272 hCoV-19/USA/NY-PV09357/2020|EPI_ISL_450045|2020-03 ···················· ···················· ····················· ···················· ···················· ····················
C 9273 hCoV-19/USA/NY-PV09358/2020|EPI_ISL_450046|2020-03 ···················· ···················· ····················· ···················· ···················· ····················
C 9274 hCoV-19/USA/NY-PV09359/2020|EPI_ISL_450047|2020-03 ···················· ···················· ····················· ···················· ···················· ····················
C 9275 hCoV-19/USA/NY-PV09360/2020|EPI_ISL_450048|2020-03 ···················· ···················· ····················· ···················· ···················· ····················

| | | | | | | | | |
|---|---|---|---|---|---|---|---|---|
| C | 9276 | hCoV-19/USA/NJ-PV09361/2020\|EPI_ISL_450049\|2020-03 | .................... | .................... | .................... | .................... | .................... | .................... |
| C | 9277 | hCoV-19/USA/NY-PV09364/2020\|EPI_ISL_450051\|2020-03 | .................... | .................... | .................... | .................... | .................... | .................... |
| C | 9278 | hCoV-19/USA/NY-PV09367/2020\|EPI_ISL_450052\|2020-03 | .................... | .................... | .................... | .................... | .................... | .................... |
| C | 9279 | hCoV-19/USA/NY-PV09368/2020\|EPI_ISL_450053\|2020-03 | .................... | .................... | .................... | .................... | .................... | .................... |
| C | 9280 | hCoV-19/USA/NY-PV09369/2020\|EPI_ISL_450054\|2020-03 | .................... | .................... | .................... | .................... | .................... | .................... |
| C | 9281 | hCoV-19/USA/NY-PV09370/2020\|EPI_ISL_450055\|2020-03 | .................... | .................... | .................... | .................... | .................... | .................... |
| C | 9282 | hCoV-19/USA/NY-PV09371/2020\|EPI_ISL_450056\|2020-03 | .................... | .................... | .................... | .................... | .................... | .................... |
| C | 9283 | hCoV-19/USA/NY-PV09374/2020\|EPI_ISL_450058\|2020-03 | .................... | .................... | .................... | .................... | .................... | .................... |
| C | 9284 | hCoV-19/USA/NY-PV09375/2020\|EPI_ISL_450059\|2020-03 | .................... | .................... | .................... | .................... | .................... | .................... |
| C | 9285 | hCoV-19/USA/NY-PV09377/2020\|EPI_ISL_450060\|2020-03 | .................... | .................... | .................... | .................... | .................... | .................... |
| C | 9286 | hCoV-19/USA/NY-PV09378/2020\|EPI_ISL_450061\|2020-03 | .................... | .................... | .................... | .................... | .................... | .................... |
| C | 9287 | hCoV-19/USA/NY-PV09379/2020\|EPI_ISL_450062\|2020-03 | .................... | .................... | .................... | .................... | .................... | .................... |
| C | 9288 | hCoV-19/USA/NY-PV09380/2020\|EPI_ISL_450063\|2020-03 | .................... | .................... | .................... | .................... | .................... | .................... |
| C | 9289 | hCoV-19/USA/NY-PV09381/2020\|EPI_ISL_450064\|2020-03 | .................... | .................... | .................... | .................... | .................... | .................... |
| C | 9290 | hCoV-19/USA/NY-PV09382/2020\|EPI_ISL_450065\|2020-03 | .................... | .................... | .................... | .................... | .................... | .................... |
| C | 9291 | hCoV-19/USA/NY-PV09383/2020\|EPI_ISL_450066\|2020-03 | .................... | .................... | .................... | .................... | .................... | .................... |
| C | 9292 | hCoV-19/Italy/LAZ-INMI1-N/2020\|EPI_ISL_451300\|2020 | .................... | .................... | .................... | .................... | .................... | .................... |
| C | 9293 | hCoV-19/Italy/LAZ-INMI2-B/2020\|EPI_ISL_451301\|2020 | .................... | .................... | .................... | .................... | .................... | .................... |
| C | 9294 | hCoV-19/Italy/LAZ-INMI2-N/2020\|EPI_ISL_451302\|2020 | .................... | .................... | .................... | .................... | .................... | .................... |
| C | 9295 | hCoV-19/Italy/LAZ-INMI5-N/2020\|EPI_ISL_451303\|2020 | .................... | .................... | .................... | .................... | .................... | .................... |
| C | 9296 | hCoV-19/Italy/LAZ-INMI11-B/2020\|EPI_ISL_451304\|202 | .................... | .................... | .................... | .................... | .................... | .................... |
| C | 9297 | hCoV-19/Italy/LOM-INMI-5314-B/2020\|EPI_ISL_451306\| | .................... | .................... | .................... | .................... | .................... | .................... |
| C | 9298 | hCoV-19/Italy/LOM-INMI-5314-N/2020\|EPI_ISL_451307\| | .................... | .................... | .................... | .................... | .................... | .................... |
| C | 9299 | hCoV-19/Italy/LOM-INMI-13075-B/2020\|EPI_ISL_451308 | .................... | .................... | .................... | .................... | .................... | .................... |
| C | 9300 | hCoV-19/Italy/LOM-INMI-13075-N/2020\|EPI_ISL_451309 | .................... | .................... | .................... | .................... | .................... | .................... |
| C | 9301 | hCoV-19/Greece/HPI-6478/2020\|EPI_ISL_451310\|2020-0 | .................... | .................... | .................... | .................... | .................... | .................... |
| C | 9302 | hCoV-19/Greece/HPI-6465/2020\|EPI_ISL_451311\|2020-0 | .................... | .................... | .................... | .................... | .................... | .................... |
| C | 9303 | hCoV-19/Sichuan/SC-GA-062/2020\|EPI_ISL_451313\|2020 | .................... | .................... | .................... | .................... | .................... | .................... |
| C | 9304 | hCoV-19/Sichuan/SC-GA-063/2020\|EPI_ISL_451314\|2020 | .................... | .................... | .................... | .................... | .................... | .................... |
| C | 9305 | hCoV-19/Sichuan/SC-GA-064/2020\|EPI_ISL_451315\|2020 | .................... | .................... | .................... | .................... | .................... | .................... |
| C | 9306 | hCoV-19/Sichuan/SC-GA-065/2020\|EPI_ISL_451316\|2020 | .................... | .................... | .................... | .................... | .................... | .................... |
| C | 9307 | hCoV-19/Sichuan/SC-LZ-079/2020\|EPI_ISL_451318\|2020 | .................... | .................... | .................... | .................... | .................... | .................... |
| C | 9308 | hCoV-19/Sichuan/SC-MY-053/2020\|EPI_ISL_451319\|2020 | .................... | .................... | .................... | .................... | .................... | .................... |
| C | 9309 | hCoV-19/Sichuan/SC-MY-054/2020\|EPI_ISL_451320\|2020 | .................... | .................... | .................... | .................... | .................... | .................... |
| C | 9310 | hCoV-19/Sichuan/SC-MY-055/2020\|EPI_ISL_451321\|2020 | .................... | .................... | .................... | .................... | .................... | .................... |
| C | 9311 | hCoV-19/Sichuan/SC-MY-056/2020\|EPI_ISL_451322\|2020 | .................... | .................... | .................... | .................... | .................... | .................... |
| C | 9312 | hCoV-19/Sichuan/SC-MY-059/2020\|EPI_ISL_451325\|2020 | .................... | .................... | .................... | .................... | .................... | .................... |
| C | 9313 | hCoV-19/Sichuan/SC-MY-060/2020\|EPI_ISL_451326\|2020 | .................... | .................... | .................... | .................... | .................... | .................... |
| C | 9314 | hCoV-19/Sichuan/SC-NC-067/2020\|EPI_ISL_451327\|2020 | .................... | .................... | .................... | .................... | .................... | .................... |
| C | 9315 | hCoV-19/Sichuan/SC-NC-068/2020\|EPI_ISL_451328\|2020 | .................... | .................... | .................... | .................... | .................... | .................... |
| C | 9316 | hCoV-19/Sichuan/SC-NC-069/2020\|EPI_ISL_451329\|2020 | .................... | .................... | .................... | .................... | .................... | .................... |
| C | 9317 | hCoV-19/Sichuan/SC-NC-071/2020\|EPI_ISL_451330\|2020 | .................... | .................... | .................... | .................... | .................... | .................... |
| C | 9318 | hCoV-19/Sichuan/SC-NC-072/2020\|EPI_ISL_451331\|2020 | .................... | .................... | .................... | .................... | .................... | .................... |
| C | 9319 | hCoV-19/Sichuan/SC-NC-075/2020\|EPI_ISL_451334\|2020 | .................... | .................... | .................... | .................... | .................... | .................... |
| C | 9320 | hCoV-19/Sichuan/SC-NC-078/2020\|EPI_ISL_451337\|2020 | .................... | .................... | .................... | .................... | .................... | .................... |
| C | 9321 | hCoV-19/Sichuan/SC-NJ-089/2020\|EPI_ISL_451338\|2020 | .................... | .................... | .................... | .................... | .................... | .................... |
| C | 9322 | hCoV-19/Sichuan/SC-WCH1-005/2020\|EPI_ISL_451376\|20 | .................... | .................... | .................... | .................... | .................... | .................... |
| C | 9323 | hCoV-19/Sichuan/SC-WCH2-006/2020\|EPI_ISL_451377\|20 | .................... | .................... | .................... | .................... | .................... | .................... |
| C | 9324 | hCoV-19/Sichuan/SC-WCH2-017/2020\|EPI_ISL_451378\|20 | .................... | .................... | .................... | .................... | .................... | .................... |
| C | 9325 | hCoV-19/Sichuan/SC-WCH-308/2020\|EPI_ISL_451379\|202 | .................... | .................... | .................... | .................... | .................... | .................... |
| C | 9326 | hCoV-19/Sichuan/SC-WCH-310/2020\|EPI_ISL_451380\|202 | .................... | .................... | .................... | .................... | .................... | .................... |
| C | 9327 | hCoV-19/Sichuan/SC-WCH-311/2020\|EPI_ISL_451381\|202 | .................... | .................... | .................... | .................... | .................... | .................... |
| C | 9328 | hCoV-19/Sichuan/SC-WCH-315/2020\|EPI_ISL_451382\|202 | .................... | .................... | .................... | .................... | .................... | .................... |
| C | 9329 | hCoV-19/Sichuan/SC-WCH3-254/2020\|EPI_ISL_451383\|20 | .................... | .................... | .................... | .................... | .................... | .................... |
| C | 9330 | hCoV-19/Sichuan/SC-WCH3-255/2020\|EPI_ISL_451384\|20 | .................... | .................... | .................... | .................... | .................... | .................... |
| C | 9331 | hCoV-19/Sichuan/SC-WCH3-259/2020\|EPI_ISL_451385\|20 | .................... | .................... | .................... | .................... | .................... | .................... |
| C | 9332 | hCoV-19/Sichuan/SC-WCH4-284/2020\|EPI_ISL_451386\|20 | .................... | .................... | .................... | .................... | .................... | .................... |
| C | 9333 | hCoV-19/Sichuan/SC-WCH4-285/2020\|EPI_ISL_451387\|20 | .................... | .................... | .................... | .................... | .................... | .................... |
| C | 9334 | hCoV-19/Sichuan/SC-WCH4-286/2020\|EPI_ISL_451388\|20 | .................... | .................... | .................... | .................... | .................... | .................... |
| C | 9335 | hCoV-19/Sichuan/SC-WCH4-287/2020\|EPI_ISL_451389\|20 | .................... | .................... | .................... | .................... | .................... | .................... |
| C | 9336 | hCoV-19/Sichuan/SC-WCH4-288/2020\|EPI_ISL_451390\|20 | .................... | .................... | .................... | .................... | .................... | .................... |
| C | 9337 | hCoV-19/Sichuan/SC-WCH4-289/2020\|EPI_ISL_451391\|20 | .................... | .................... | .................... | .................... | .................... | .................... |
| C | 9338 | hCoV-19/Sichuan/SC-WCH4-290/2020\|EPI_ISL_451392\|20 | .................... | .................... | .................... | .................... | .................... | .................... |
| C | 9339 | hCoV-19/Sichuan/SC-WCH4-291/2020\|EPI_ISL_451393\|20 | .................... | .................... | .................... | .................... | .................... | .................... |
| C | 9340 | hCoV-19/Sichuan/SC-WCH4-293/2020\|EPI_ISL_451394\|20 | .................... | .................... | .................... | .................... | .................... | .................... |
| C | 9341 | hCoV-19/Sichuan/SC-WCH4-301/2020\|EPI_ISL_451395\|20 | .................... | .................... | .................... | .................... | .................... | .................... |
| C | 9343 | hCoV-19/Spain/PV-IBV-000941/2020\|EPI_ISL_452631\|20 | .................... | .................... | .................... | .................... | .................... | .................... |
| C | 9349 | hCoV-19/Spain/PV-IBV-000935/2020\|EPI_ISL_452637\|20 | .................... | .................... | .................... | .................... | .................... | .................... |
| C | 9359 | hCoV-19/Spain/PV-IBV-000924/2020\|EPI_ISL_452647\|20 | .................... | .................... | .................... | .................... | .................... | .................... |
| C | 9363 | hCoV-19/Spain/PV-IBV-000918/2020\|EPI_ISL_452651\|20 | .................... | .................... | .................... | .................... | .................... | .................... |
| C | 9377 | hCoV-19/Spain/PV-IBV-000892/2020\|EPI_ISL_452671\|20 | .................... | .................... | .................... | .................... | .................... | .................... |
| C | 9379 | hCoV-19/Portugal/PT0247/2020\|EPI_ISL_453961\|2020-0 | .................... | .................... | .................... | .................... | .................... | .................... |
| C | 9380 | hCoV-19/Portugal/PT0248/2020\|EPI_ISL_453962\|2020-0 | .................... | .................... | .................... | .................... | .................... | .................... |
| C | 9381 | hCoV-19/Portugal/PT0249/2020\|EPI_ISL_453963\|2020-0 | .................... | .................... | .................... | .................... | .................... | .................... |
| C | 9382 | hCoV-19/Portugal/PT0250/2020\|EPI_ISL_453964\|2020-0 | .................... | .................... | .................... | .................... | .................... | .................... |
| C | 9383 | hCoV-19/Portugal/PT0251/2020\|EPI_ISL_453965\|2020-0 | .................... | .................... | .................... | .................... | .................... | .................... |
| C | 9384 | hCoV-19/Portugal/PT0252/2020\|EPI_ISL_453966\|2020-0 | .................... | .................... | .................... | .................... | .................... | .................... |
| C | 9385 | hCoV-19/Portugal/PT0253/2020\|EPI_ISL_453967\|2020-0 | .................... | .................... | .................... | .................... | .................... | .................... |

| | | | | | | | | | |
|---|---|---|---|---|---|---|---|---|---|
| C | 9386 | hCoV-19/Portugal/PT0255/2020\|EPI_ISL_453969\|2020-0 | .................... | .................... | .................... | .................... | .................... | .................... | |
| C | 9387 | hCoV-19/Portugal/PT0256/2020\|EPI_ISL_453970\|2020-0 | .................... | .................... | .................... | .................... | .................... | .................... | |
| C | 9388 | hCoV-19/Portugal/PT0257/2020\|EPI_ISL_453971\|2020-0 | .................... | .................... | .................... | .................... | .................... | .................... | |
| C | 9389 | hCoV-19/Portugal/PT0258/2020\|EPI_ISL_453972\|2020-0 | .................... | .................... | .................... | .................... | .................... | .................... | |
| C | 9390 | hCoV-19/Portugal/PT0259/2020\|EPI_ISL_453973\|2020-0 | .................... | .................... | .................... | .................... | .................... | .................... | |
| C | 9391 | hCoV-19/Portugal/PT0260/2020\|EPI_ISL_453974\|2020-0 | .................... | .................... | .................... | .................... | .................... | .................... | |
| C | 9392 | hCoV-19/Portugal/PT0261/2020\|EPI_ISL_453975\|2020-0 | .................... | .................... | .................... | .................... | .................... | .................... | |
| C | 9393 | hCoV-19/Portugal/PT0262/2020\|EPI_ISL_453976\|2020-0 | .................... | .................... | .................... | .................... | .................... | .................... | |
| C | 9394 | hCoV-19/Portugal/PT0263/2020\|EPI_ISL_453977\|2020-0 | .................... | .................... | .................... | .................... | .................... | .................... | |
| C | 9395 | hCoV-19/Portugal/PT0265/2020\|EPI_ISL_453979\|2020-0 | .................... | .................... | .................... | .................... | .................... | .................... | |
| C | 9396 | hCoV-19/Portugal/PT0267/2020\|EPI_ISL_453981\|2020-0 | .................... | .................... | .................... | .................... | .................... | .................... | |
| C | 9397 | hCoV-19/Portugal/PT0268/2020\|EPI_ISL_453982\|2020-0 | .................... | .................... | .................... | .................... | .................... | .................... | |
| C | 9398 | hCoV-19/Portugal/PT0270/2020\|EPI_ISL_453984\|2020-0 | .................... | .................... | .................... | .................... | .................... | .................... | |
| C | 9399 | hCoV-19/Portugal/PT0272/2020\|EPI_ISL_453986\|2020-0 | .................... | .................... | .................... | .................... | .................... | .................... | |
| C | 9400 | hCoV-19/Portugal/PT0273/2020\|EPI_ISL_453987\|2020-0 | .................... | .................... | .................... | .................... | .................... | .................... | |
| C | 9401 | hCoV-19/Portugal/PT0274/2020\|EPI_ISL_453988\|2020-0 | .................... | .................... | .................... | .................... | .................... | .................... | |
| C | 9402 | hCoV-19/Portugal/PT0275/2020\|EPI_ISL_453989\|2020-0 | .................... | .................... | .................... | .................... | .................... | .................... | |
| C | 9403 | hCoV-19/Portugal/PT0276/2020\|EPI_ISL_453990\|2020-0 | .................... | .................... | .................... | .................... | .................... | .................... | |
| C | 9404 | hCoV-19/Portugal/PT0278/2020\|EPI_ISL_453992\|2020-0 | .................... | .................... | .................... | .................... | .................... | .................... | |
| C | 9405 | hCoV-19/Portugal/PT0279/2020\|EPI_ISL_453993\|2020-0 | .................... | .................... | .................... | .................... | .................... | .................... | |
| C | 9406 | hCoV-19/Portugal/PT0282/2020\|EPI_ISL_453996\|2020-0 | .................... | .................... | .................... | .................... | .................... | .................... | |
| C | 9407 | hCoV-19/Portugal/PT0283a/2020\|EPI_ISL_453997\|2020- | .................... | .................... | .................... | .................... | .................... | .................... | |
| C | 9408 | hCoV-19/Portugal/PT0283b/2020\|EPI_ISL_453998\|2020- | .................... | .................... | .................... | .................... | .................... | .................... | |
| C | 9409 | hCoV-19/Portugal/PT0284/2020\|EPI_ISL_453999\|2020-0 | .................... | .................... | .................... | .................... | .................... | .................... | |
| C | 9410 | hCoV-19/England/NORW-EBCF8/2020\|EPI_ISL_457500\|202 | .................... | .................... | .................... | .................... | .................... | .................... | |
| C | 9411 | hCoV-19/England/NORW-EBD04/2020\|EPI_ISL_457501\|202 | .................... | ...................A | .................... | .................... | .................... | .................... | 1 |
| C | 9412 | hCoV-19/England/NORW-EBD31/2020\|EPI_ISL_457503\|202 | .................... | .................... | .................... | .................... | .................... | .................... | |
| C | 9413 | hCoV-19/England/NORW-EBD40/2020\|EPI_ISL_457504\|202 | .................... | .................... | .................... | .................... | .................... | .................... | |
| C | 9414 | hCoV-19/England/NORW-EBD5F/2020\|EPI_ISL_457505\|202 | .................... | .................... | .................... | .................... | .................... | .................... | |
| C | 9415 | hCoV-19/England/NORW-EBD7D/2020\|EPI_ISL_457506\|202 | .................... | .................... | .................... | .................... | .................... | .................... | |
| C | 9416 | hCoV-19/England/NORW-EBD8C/2020\|EPI_ISL_457507\|202 | .................... | .................... | .................... | .................... | .................... | .................... | |
| C | 9417 | hCoV-19/England/NORW-EBDB9/2020\|EPI_ISL_457509\|202 | .................... | .................... | .................... | .................... | .................... | .................... | |
| C | 9418 | hCoV-19/England/NORW-EBDC8/2020\|EPI_ISL_457510\|202 | .................... | .................... | .................... | .................... | .................... | .................... | |
| C | 9419 | hCoV-19/England/NORW-EBDE6/2020\|EPI_ISL_457511\|202 | .................... | .................... | .................... | .................... | .................... | .................... | |
| C | 9420 | hCoV-19/England/NORW-EBDF5/2020\|EPI_ISL_457512\|202 | .................... | .................... | .................... | .................... | .................... | .................... | |
| C | 9421 | hCoV-19/England/NORW-EBE01/2020\|EPI_ISL_457513\|202 | .................... | .................... | .................... | .................... | .................... | .................... | |
| C | 9422 | hCoV-19/England/NORW-EBE10/2020\|EPI_ISL_457514\|202 | .................... | .................... | .................... | .................... | .................... | .................... | |
| C | 9423 | hCoV-19/England/NORW-EBE2F/2020\|EPI_ISL_457515\|202 | .................... | .................... | .................... | .................... | .................... | .................... | |
| C | 9424 | hCoV-19/England/NORW-EBE3E/2020\|EPI_ISL_457516\|202 | .................... | .................... | .................... | .................... | .................... | .................... | |
| C | 9425 | hCoV-19/England/NORW-EBE6B/2020\|EPI_ISL_457517\|202 | .................... | .................... | .................... | .................... | .................... | .................... | |
| C | 9426 | hCoV-19/England/NORW-EBE89/2020\|EPI_ISL_457518\|202 | .................... | .................... | .................... | .................... | .................... | .................... | |
| C | 9427 | hCoV-19/England/NORW-EC35C/2020\|EPI_ISL_457520\|202 | .................... | .................... | .................... | .................... | .................... | .................... | |
| C | 9428 | hCoV-19/England/NORW-EC389/2020\|EPI_ISL_457521\|202 | .................... | .................... | .................... | .................... | .................... | .................... | |
| C | 9429 | hCoV-19/England/NORW-EC40E/2020\|EPI_ISL_457523\|202 | .................... | .................... | .................... | .................... | .................... | .................... | |
| C | 9430 | hCoV-19/England/NORW-EC41D/2020\|EPI_ISL_457524\|202 | .................... | .................... | .................... | .................... | .................... | .................... | |
| C | 9431 | hCoV-19/England/BRIS-128B1F/2020\|EPI_ISL_458814\|20 | .................... | .................... | .................... | .................... | .................... | .................... | |
| C | 9432 | hCoV-19/England/BRIS-12948C/2020\|EPI_ISL_458815\|20 | .................... | .................... | .................... | .................... | .................... | .................... | |
| C | 9433 | hCoV-19/England/BRIS-12920A/2020\|EPI_ISL_458816\|20 | .................... | .................... | .................... | .................... | .................... | .................... | |
| C | 9434 | hCoV-19/England/BRIS-129677/2020\|EPI_ISL_458817\|20 | .................... | .................... | .................... | .................... | .................... | .................... | |
| C | 9435 | hCoV-19/England/BRIS-128DDD/2020\|EPI_ISL_458819\|20 | .................... | .................... | .................... | .................... | .................... | .................... | |
| C | 9436 | hCoV-19/England/BRIS-1293E9/2020\|EPI_ISL_458820\|20 | .................... | .................... | .................... | .................... | .................... | .................... | |
| C | 9437 | hCoV-19/England/BRIS-129738/2020\|EPI_ISL_458821\|20 | .................... | .................... | .................... | .................... | .................... | .................... | |
| C | 9438 | hCoV-19/England/BRIS-129176/2020\|EPI_ISL_458822\|20 | .................... | .................... | .................... | .................... | .................... | .................... | |
| C | 9439 | hCoV-19/England/BRIS-12872A/2020\|EPI_ISL_458824\|20 | .................... | .................... | .................... | .................... | .................... | .................... | |
| C | 9440 | hCoV-19/England/BRIS-1297CF/2020\|EPI_ISL_458825\|20 | .................... | .................... | .................... | .................... | .................... | .................... | |
| C | 9441 | hCoV-19/England/BRIS-129264/2020\|EPI_ISL_458826\|20 | .................... | .................... | .................... | .................... | .................... | .................... | |
| C | 9442 | hCoV-19/England/BRIS-1290B5/2020\|EPI_ISL_458827\|20 | .................... | .................... | .................... | .................... | .................... | .................... | |
| C | 9443 | hCoV-19/England/BRIS-128E9E/2020\|EPI_ISL_458829\|20 | .................... | .................... | .................... | .................... | .................... | .................... | |
| C | 9444 | hCoV-19/England/BRIS-129783/2020\|EPI_ISL_458830\|20 | .................... | .................... | .................... | .................... | .................... | .................... | |
| C | 9445 | hCoV-19/England/BRIS-128DEC/2020\|EPI_ISL_458831\|20 | .................... | .................... | .................... | .................... | .................... | .................... | |
| C | 9446 | hCoV-19/England/BRIS-128A12/2020\|EPI_ISL_458832\|20 | .................... | .................... | .................... | .................... | .................... | .................... | |
| C | 9447 | hCoV-19/England/BRIS-128EDA/2020\|EPI_ISL_458833\|20 | .................... | ............A....... | .................... | .................... | .................... | .................... | |
| C | 9448 | hCoV-19/England/BRIS-128F7D/2020\|EPI_ISL_458835\|20 | .................... | .................... | .................... | .................... | .................... | .................... | |
| C | 9449 | hCoV-19/England/BRIS-129510/2020\|EPI_ISL_458836\|20 | .................... | .................... | .................... | .................... | .................... | .................... | |
| C | 9450 | hCoV-19/England/BRIS-128DBF/2020\|EPI_ISL_458838\|20 | .................... | .................... | .................... | .................... | .................... | .................... | |
| C | 9451 | hCoV-19/England/BRIS-128F6E/2020\|EPI_ISL_458839\|20 | .................... | .................... | .................... | .................... | .................... | .................... | |
| C | 9452 | hCoV-19/England/BRIS-129307/2020\|EPI_ISL_458840\|20 | .................... | .................... | .................... | .................... | .................... | .................... | |
| C | 9453 | hCoV-19/England/BRIS-12871B/2020\|EPI_ISL_458842\|20 | .................... | .................... | .................... | .................... | .................... | .................... | |
| C | 9454 | hCoV-19/England/BRIS-128D55/2020\|EPI_ISL_458844\|20 | .................... | .................... | .................... | .................... | .................... | .................... | |
| C | 9455 | hCoV-19/England/BRIS-128EBC/2020\|EPI_ISL_458847\|20 | .................... | .................... | .................... | .................... | .................... | .................... | |
| C | 9456 | hCoV-19/England/BRIS-128784/2020\|EPI_ISL_458848\|20 | .................... | .................... | .................... | .................... | .................... | .................... | |
| C | 9457 | hCoV-19/England/BRIS-1296B3/2020\|EPI_ISL_458851\|20 | .................... | .................... | .................... | .................... | .................... | .................... | |
| C | 9458 | hCoV-19/England/BRIS-12904C/2020\|EPI_ISL_458854\|20 | .................... | .................... | .................... | .................... | .................... | .................... | |
| C | 9459 | hCoV-19/England/SHEF-C6886/2020\|EPI_ISL_462025\|202 | .................... | .................... | .................... | .................... | .................... | .................... | |
| C | 9460 | hCoV-19/England/SHEF-C721E/2020\|EPI_ISL_462029\|202 | .................... | .................... | .................... | .................... | .................... | .................... | |
| C | 9461 | hCoV-19/England/SHEF-C7409/2020\|EPI_ISL_462030\|202 | .................... | .................... | .................... | .................... | .................... | .................... | |
| C | 9462 | hCoV-19/England/SHEF-C7885/2020\|EPI_ISL_462032\|202 | .................... | .................... | .................... | .................... | .................... | .................... | |
| C | 9463 | hCoV-19/England/SHEF-C7F26/2020\|EPI_ISL_462034\|202 | .................... | .................... | .................... | .................... | .................... | .................... | |

| | | | | | | | | | |
|---|---|---|---|---|---|---|---|---|---|
| C | 9464 | hCoV-19/England/SHEF-C83FC/2020\|EPI_ISL_462036\|202 | ···················· | ···················· | ···················· | ···················· | ···················· | ···················· | |
| C | 9465 | hCoV-19/England/SHEF-C92D0/2020\|EPI_ISL_462038\|202 | ···················· | ···················· | ···················· | ···················· | ···················· | ···················· | |
| C | 9466 | hCoV-19/England/SHEF-C9610/2020\|EPI_ISL_462039\|202 | ···················· | ···················· | ···················· | ···················· | ···················· | ···················· | |
| C | 9467 | hCoV-19/England/SHEF-C98DE/2020\|EPI_ISL_462041\|202 | ···················· | ···················· | ···················· | ···················· | ···················· | ···················· | |
| C | 9468 | hCoV-19/England/SHEF-C9F9D/2020\|EPI_ISL_462043\|202 | ···················· | ···················· | ···················· | ···················· | ···················· | ···················· | |
| C | 9469 | hCoV-19/England/SHEF-CA58B/2020\|EPI_ISL_462044\|202 | ···················· | ···················· | ···················· | ···················· | ···················· | ···················· | |
| C | 9470 | hCoV-19/England/SHEF-CA6A6/2020\|EPI_ISL_462045\|202 | ···················· | ···················· | ···················· | ···················· | ···················· | ···················· | |
| C | 9471 | hCoV-19/England/SHEF-D16E8/2020\|EPI_ISL_462079\|202 | ···················· | ···················· | ···················· | ···················· | ···················· | ···················· | |
| C | 9472 | hCoV-19/England/SHEF-D1EE0/2020\|EPI_ISL_462081\|202 | ···················· | ···················· | ···················· | ···················· | ···················· | ···················· | |
| C | 9473 | hCoV-19/England/NORW-E739D/2020\|EPI_ISL_449030\|202 | ···················· | ···················· | ···················· | ···················· | ···················· | ···················· | |
| C | 9474 | hCoV-19/England/NORW-E73BB/2020\|EPI_ISL_449031\|202 | ···················· | ···················· | ···················· | ···················· | ···················· | ···················· | |
| C | 9475 | hCoV-19/England/NORW-E73CA/2020\|EPI_ISL_449032\|202 | ···················· | ···················· | ···················· | ···················· | ···················· | ···················· | |
| C | 9476 | hCoV-19/England/NORW-E7421/2020\|EPI_ISL_449034\|202 | ···················· | ···················· | ···················· | ···················· | ···················· | ···················· | |
| C | 9477 | hCoV-19/England/NORW-E7430/2020\|EPI_ISL_449035\|202 | ···················· | ···················· | ···················· | ···················· | ···················· | ···················· | |
| C | 9478 | hCoV-19/England/NORW-E744F/2020\|EPI_ISL_449036\|202 | ···················· | ···················· | ···················· | ···················· | ···················· | ···················· | |
| C | 9479 | hCoV-19/England/NORW-E747C/2020\|EPI_ISL_449037\|202 | ···················· | ···················· | ···················· | ···················· | ···················· | ···················· | |
| C | 9480 | hCoV-19/England/NORW-E748B/2020\|EPI_ISL_449038\|202 | ···················· | ···················· | ···················· | ···················· | ···················· | ···················· | |
| C | 9481 | hCoV-19/England/NORW-E749A/2020\|EPI_ISL_449039\|202 | ···················· | ···················· | ···················· | ···················· | ···················· | ···················· | |
| C | 9482 | hCoV-19/England/NORW-E74A9/2020\|EPI_ISL_449040\|202 | ···················· | ···················· | ···················· | ···················· | ···················· | ···················· | |
| C | 9483 | hCoV-19/England/NORW-E74B8/2020\|EPI_ISL_449041\|202 | ···················· | ···················· | ···················· | ···················· | ···················· | ···················· | |
| C | 9484 | hCoV-19/England/NORW-E74E5/2020\|EPI_ISL_449042\|202 | ···················· | ···················· | ···················· | ···················· | ···················· | ···················· | |
| C | 9485 | hCoV-19/England/NORW-E74F4/2020\|EPI_ISL_449043\|202 | ···················· | ···················· | ···················· | ···················· | ···················· | ···················· | |
| C | 9486 | hCoV-19/England/NORW-E7500/2020\|EPI_ISL_449044\|202 | ···················· | ···················· | ···················· | ···················· | ···················· | ···················· | |
| C | 9487 | hCoV-19/England/NORW-E751F/2020\|EPI_ISL_449045\|202 | ···················· | ···················· | ···················· | ···················· | ···················· | ···················· | |
| C | 9488 | hCoV-19/England/NORW-E753D/2020\|EPI_ISL_449046\|202 | ···················· | ···················· | ···················· | ···················· | ···················· | ···················· | |
| C | 9489 | hCoV-19/England/NORW-E754C/2020\|EPI_ISL_449047\|202 | ···················· | ···················· | ···················· | ···················· | NNN················· | ···················· | 1 |
| C | 9490 | hCoV-19/England/NORW-E755B/2020\|EPI_ISL_449048\|202 | ···················· | ···················· | ···················· | ···················· | ···················· | ···················· | |
| C | 9491 | hCoV-19/England/NORW-E756A/2020\|EPI_ISL_449049\|202 | ···················· | ···················· | ···················· | ···················· | ···················· | ···················· | |
| C | 9492 | hCoV-19/England/NORW-E7676/2020\|EPI_ISL_449060\|202 | ···················· | ···················· | ···················· | ···················· | ···················· | ···················· | |
| C | 9493 | hCoV-19/England/NORW-E7685/2020\|EPI_ISL_449061\|202 | ···················· | ···················· | ···················· | ···················· | ···················· | ···················· | |
| C | 9494 | hCoV-19/England/NORW-E7694/2020\|EPI_ISL_449062\|202 | ···················· | ···················· | ···················· | ···················· | ···················· | ···················· | |
| C | 9495 | hCoV-19/England/NORW-E76B2/2020\|EPI_ISL_449063\|202 | ···················· | ···················· | ···················· | ···················· | ···················· | ···················· | |
| C | 9496 | hCoV-19/England/NORW-E76FE/2020\|EPI_ISL_449065\|202 | ···················· | ···················· | ···················· | ···················· | ···················· | ···················· | |
| C | 9497 | hCoV-19/England/NORW-E7737/2020\|EPI_ISL_449066\|202 | ··················T· | ···················· | ···················· | ···················· | ···················· | ···················· | 1 |
| C | 9499 | hCoV-19/England/NORW-E7782/2020\|EPI_ISL_449069\|202 | ···················· | ···················· | ···················· | ···················· | ···················· | ···················· | |
| C | 9500 | hCoV-19/USA/NY-PV09323/2020\|EPI_ISL_450012\|2020-03 | ···················· | ···················· | ···················· | ···················· | ···················· | ···················· | |
| C | 9501 | hCoV-19/USA/NY-PV09324/2020\|EPI_ISL_450013\|2020-03 | ···················· | ·····A·············· | ···················· | ···················· | ···················· | ···················· | |
| C | 9502 | hCoV-19/USA/NY-PV09325/2020\|EPI_ISL_450014\|2020-03 | ···················· | ···················· | ···················· | ···················· | ···················· | ···················· | |
| C | 9503 | hCoV-19/USA/NY-PV09326/2020\|EPI_ISL_450015\|2020-03 | ···················· | ···················· | ···················· | ···················· | ···················· | ···················· | |
| C | 9504 | hCoV-19/USA/NY-PV09327/2020\|EPI_ISL_450016\|2020-03 | ···················· | ···················· | ···················· | ···················· | ···················· | ···················· | |
| C | 9505 | hCoV-19/USA/NY-PV09328/2020\|EPI_ISL_450017\|2020-03 | ···················· | ············A······· | ···················· | ···················· | ···················· | ···················· | |
| C | 9506 | hCoV-19/USA/NY-PV09329/2020\|EPI_ISL_450018\|2020-03 | ···················· | ···················· | ···················· | ···················· | ···················· | ···················· | |
| C | 9507 | hCoV-19/USA/NY-PV09330/2020\|EPI_ISL_450019\|2020-03 | ···················· | ···················· | ···················· | ···················· | ···················· | ···················· | |
| C | 9508 | hCoV-19/USA/NY-PV09331/2020\|EPI_ISL_450020\|2020-03 | ···················· | ···················· | ···················· | ···················· | ···················· | ···················· | |
| C | 9509 | hCoV-19/USA/NY-PV09333/2020\|EPI_ISL_450021\|2020-03 | ···················· | ···················· | ···················· | ···················· | ···················· | ···················· | |
| C | 9510 | hCoV-19/USA/NY-PV09334/2020\|EPI_ISL_450022\|2020-03 | ···················· | ···················· | ···················· | ···················· | ···················· | ···················· | |
| C | 9511 | hCoV-19/USA/NY-PV09335/2020\|EPI_ISL_450023\|2020-03 | ···················· | ···················· | ···················· | ···················· | ···················· | ···················· | |
| C | 9512 | hCoV-19/USA/NY-PV09336/2020\|EPI_ISL_450024\|2020-03 | ···················· | ···················· | ···················· | ···················· | ···················· | ···················· | |
| C | 9513 | hCoV-19/USA/NY-PV09337/2020\|EPI_ISL_450025\|2020-03 | ···················· | ···················· | ···················· | ···················· | ···················· | ···················· | |
| C | 9514 | hCoV-19/USA/NY-PV09338/2020\|EPI_ISL_450026\|2020-03 | ···················· | ···················· | ···················· | ···················· | ···················· | ···················· | |
| C | 9515 | hCoV-19/USA/NY-PV09339/2020\|EPI_ISL_450027\|2020-03 | ···················· | ···················· | ···················· | ···················· | ···················· | ···················· | |
| C | 9516 | hCoV-19/USA/NY-PV09340/2020\|EPI_ISL_450028\|2020-03 | ···················· | ···················· | ···················· | ···················· | ···················· | ···················· | |
| C | 9517 | hCoV-19/USA/NY-PV09341/2020\|EPI_ISL_450029\|2020-03 | ···················· | ···················· | ···················· | ···················· | ···················· | ···················· | |
| C | 9518 | hCoV-19/USA/NY-PV09342/2020\|EPI_ISL_450030\|2020-03 | ···················· | ···················· | ···················· | ···················· | ···················· | ···················· | |
| C | 9519 | hCoV-19/USA/NY-PV09343/2020\|EPI_ISL_450031\|2020-03 | ···················· | ···················· | ···················· | ···················· | ···················· | ···················· | |
| C | 9520 | hCoV-19/USA/NY-PV09344/2020\|EPI_ISL_450032\|2020-03 | ···················· | ···················· | ···················· | ···················· | ···················· | ···················· | |
| C | 9521 | hCoV-19/USA/NY-PV09345/2020\|EPI_ISL_450033\|2020-03 | ···················· | ···················· | ···················· | ···················· | ···················· | ···················· | |
| C | 9522 | hCoV-19/USA/NY-PV09346/2020\|EPI_ISL_450034\|2020-03 | ···················· | ···················· | ···················· | ···················· | ···················· | ···················· | |
| C | 9523 | hCoV-19/USA/NY-PV09347/2020\|EPI_ISL_450035\|2020-03 | ···················· | ···················· | ···················· | ···················· | ···················· | ···················· | |
| C | 9524 | hCoV-19/USA/NY-PV09348/2020\|EPI_ISL_450036\|2020-03 | ···················· | ···················· | ···················· | ···················· | ···················· | ···················· | |
| C | 9525 | hCoV-19/USA/NY-PV09349/2020\|EPI_ISL_450037\|2020-03 | ···················· | ···················· | ···················· | ···················· | ···················· | ···················· | |
| C | 9526 | hCoV-19/USA/NY-PV09350/2020\|EPI_ISL_450038\|2020-03 | ···················· | ···················· | ···················· | ···················· | ···················· | ···················· | |
| C | 9527 | hCoV-19/USA/NY-PV09351/2020\|EPI_ISL_450039\|2020-03 | ···················· | ···················· | ···················· | ···················· | ···················· | ···················· | |
| C | 9528 | hCoV-19/USA/NY-PV09352/2020\|EPI_ISL_450040\|2020-03 | ···················· | ···················· | ···················· | ···················· | ···················· | ···················· | |
| C | 9529 | hCoV-19/USA/NY-PV09353/2020\|EPI_ISL_450041\|2020-03 | ···················· | ···················· | ···················· | ···················· | ···················· | ···················· | |
| C | 9530 | hCoV-19/USA/NY-PV09354/2020\|EPI_ISL_450042\|2020-03 | ···················· | ···················· | ···················· | ···················· | ···················· | ···················· | |
| C | 9531 | hCoV-19/USA/NY-PV09355/2020\|EPI_ISL_450043\|2020-03 | ···················· | ···················· | ···················· | ···················· | ···················· | ···················· | |
| C | 9532 | hCoV-19/USA/NY-PV09356/2020\|EPI_ISL_450044\|2020-03 | ···················· | ···················· | ···················· | ···················· | ···················· | ···················· | |
| C | 9533 | hCoV-19/USA/NY-PV09362/2020\|EPI_ISL_450050\|2020-03 | ···················· | ···················· | ···················· | ···················· | ···················· | ···················· | |
| C | 9534 | hCoV-19/Sichuan/SC-NJ-095/2020\|EPI_ISL_451344\|2020 | ···················· | ···················· | ···················· | ···················· | ···················· | ···················· | |
| C | 9535 | hCoV-19/Sichuan/SC-PHCC1-022/2020\|EPI_ISL_451345\|2 | ···················· | ···················· | ···················· | ···················· | ···················· | ···················· | |
| C | 9536 | hCoV-19/Sichuan/SC-PHCC1-025/2020\|EPI_ISL_451346\|2 | ···················· | ···················· | ···················· | ···················· | ···················· | ···················· | |
| C | 9537 | hCoV-19/Sichuan/SC-PHCC1-027/2020\|EPI_ISL_451348\|2 | ···················· | ···················· | ···················· | ···················· | ···················· | ···················· | |
| C | 9538 | hCoV-19/Sichuan/SC-PHCC1-032/2020\|EPI_ISL_451353\|2 | ···················· | ···················· | ···················· | ···················· | ···················· | ···················· | |
| C | 9539 | hCoV-19/Sichuan/SC-PHCC1-033/2020\|EPI_ISL_451354\|2 | ···················· | ···················· | ···················· | ···················· | ···················· | ···················· | |
| C | 9540 | hCoV-19/Sichuan/SC-PHCC1-035/2020\|EPI_ISL_451356\|2 | ···················· | ···················· | ···················· | ···················· | ···················· | ···················· | |
| C | 9541 | hCoV-19/Sichuan/SC-PHCC1-036/2020\|EPI_ISL_451357\|2 | ···················· | ···················· | ···················· | ···················· | ···················· | ···················· | |
| C | 9542 | hCoV-19/Sichuan/SC-PHCC2-039/2020\|EPI_ISL_451359\|2 | ···················· | ···················· | ···················· | ···················· | ···················· | ···················· | |

| | | | | | | | | |
|---|---|---|---|---|---|---|---|---|
| C | 9543 hCoV-19/Sichuan/SC-PHCC2-040/2020\|EPI_ISL_451360\|2 | ···················· | ···················· | ···················· | ···················· | ···················· | ···················· | |
| C | 9544 hCoV-19/Sichuan/SC-PHCC2-045/2020\|EPI_ISL_451365\|2 | ···················· | ···················· | ···················· | ···················· | ···················· | ···················· | |
| C | 9545 hCoV-19/Sichuan/SC-PHCC2-049/2020\|EPI_ISL_451369\|2 | ···················· | ··········T········· | ···················· | ···················· | ···················· | ···················· | |
| C | 9546 hCoV-19/Sichuan/SC-PHCC2-050/2020\|EPI_ISL_451370\|2 | ···················· | ···················· | ···················· | ···················· | ···················· | ···················· | |
| C | 9547 hCoV-19/Sichuan/SC-PHCC2-051/2020\|EPI_ISL_451371\|2 | ···················· | ···················· | ···················· | ···················· | ···················· | ···················· | |
| C | 9548 hCoV-19/Sichuan/SC-WCH-082/2020\|EPI_ISL_451374\|202 | ···················· | ···················· | ···················· | ···················· | ···················· | ···················· | |
| C | 9550 hCoV-19/Spain/AN-IBV-001085/2020\|EPI_ISL_452601\|20 | ···················· | ···················· | ···················· | ···················· | ···················· | ···················· | |
| C | 9554 hCoV-19/Spain/AN-IBV-001078/2020\|EPI_ISL_452606\|20 | ···················· | ···················· | ···················· | ···················· | ···················· | ···················· | |
| C | 9561 hCoV-19/Spain/PV-IBV-000955/2020\|EPI_ISL_452620\|20 | ···················· | ···················· | ···················· | ···················· | ···················· | ···················· | |
| C | 9570 hCoV-19/Spain/PV-IBV-000882/2020\|EPI_ISL_452677\|20 | ···················· | ···················· | ···················· | ···················· | ···················· | ···················· | |
| C | 9575 hCoV-19/Spain/PV-IBV-000875/2020\|EPI_ISL_452682\|20 | ···················· | ···················· | ···················· | ···················· | ···················· | ···················· | |
| C | 9576 hCoV-19/Spain/PV-IBV-000874/2020\|EPI_ISL_452683\|20 | ···················· | ···················· | ···················· | ···················· | ···················· | ···················· | |
| C | 9577 hCoV-19/Spain/PV-IBV-000873/2020\|EPI_ISL_452684\|20 | ···················· | ···················· | ···················· | ···················· | ···················· | ···················· | |
| C | 9589 hCoV-19/Portugal/PT0185/2020\|EPI_ISL_453901\|2020-0 | ···················· | ···················· | ···················· | ···················· | ···················· | ···················· | |
| C | 9590 hCoV-19/Portugal/PT0190/2020\|EPI_ISL_453906\|2020-0 | ···················· | ···················· | ···················· | ···················· | ···················· | ···················· | |
| C | 9591 hCoV-19/Portugal/PT0195/2020\|EPI_ISL_453911\|2020-0 | ···················· | ···················· | ···················· | ···················· | ···················· | ···················· | |
| C | 9592 hCoV-19/Portugal/PT0196/2020\|EPI_ISL_453912\|2020-0 | ···················· | ···················· | ···················· | ···················· | ···················· | ···················· | |
| C | 9593 hCoV-19/Portugal/PT0197/2020\|EPI_ISL_453913\|2020-0 | ···················· | ···················· | ···················· | ···················· | ···················· | ···················· | |
| C | 9594 hCoV-19/Portugal/PT0198/2020\|EPI_ISL_453914\|2020-0 | ···················· | ···················· | ···················· | ···················· | ···················· | ···················· | |
| C | 9595 hCoV-19/Portugal/PT0199/2020\|EPI_ISL_453915\|2020-0 | ···················· | ···················· | ···················· | ···················· | ···················· | ···················· | |
| C | 9596 hCoV-19/Portugal/PT0200/2020\|EPI_ISL_453916\|2020-0 | ···················· | ···················· | ···················· | ···················· | ···················· | ···················· | |
| C | 9597 hCoV-19/Portugal/PT0203/2020\|EPI_ISL_453917\|2020-0 | ···················· | ···················· | ···················· | ···················· | ···················· | ···················· | |
| C | 9598 hCoV-19/Portugal/PT0208/2020\|EPI_ISL_453922\|2020-0 | ···················· | ···················· | ···················· | ···················· | ···················· | ···················· | |
| C | 9599 hCoV-19/Portugal/PT0210/2020\|EPI_ISL_453924\|2020-0 | ···················· | ···················· | ···················· | ···················· | ···················· | ···················· | |
| C | 9600 hCoV-19/Portugal/PT0211/2020\|EPI_ISL_453925\|2020-0 | ···················· | ···················· | ···················· | ···················· | ···················· | ···················· | |
| C | 9601 hCoV-19/Portugal/PT0213/2020\|EPI_ISL_453927\|2020-0 | ···················· | ···················· | ···················· | ···················· | ···················· | ···················· | |
| C | 9602 hCoV-19/Portugal/PT0215/2020\|EPI_ISL_453929\|2020-0 | ···················· | ···················· | ···················· | ···················· | ···················· | ···················· | |
| C | 9603 hCoV-19/Portugal/PT0217/2020\|EPI_ISL_453931\|2020-0 | ···················· | ···················· | ···················· | ···················· | ···················· | ···················· | |
| C | 9604 hCoV-19/Portugal/PT0219/2020\|EPI_ISL_453933\|2020-0 | ···················· | ···················· | ···················· | ···················· | ···················· | ···················· | |
| C | 9605 hCoV-19/Portugal/PT0225/2020\|EPI_ISL_453939\|2020-0 | ···················· | ···················· | ···················· | ···················· | ···················· | ···················· | |
| C | 9606 hCoV-19/Portugal/PT0228/2020\|EPI_ISL_453942\|2020-0 | ···················· | ···················· | ···················· | ···················· | ···················· | ···················· | |
| C | 9607 hCoV-19/Portugal/PT0229/2020\|EPI_ISL_453943\|2020-0 | ···················· | ···················· | ···················· | ···················· | ···················· | ···················· | |
| C | 9608 hCoV-19/Portugal/PT0230/2020\|EPI_ISL_453944\|2020-0 | ···················· | ···················· | ···················· | ···················· | ···················G | ···················· | 1 |
| C | 9609 hCoV-19/Portugal/PT0232/2020\|EPI_ISL_453946\|2020-0 | ···················· | ···················· | ···················· | ···················· | ···················· | ···················· | |
| C | 9610 hCoV-19/Portugal/PT0233/2020\|EPI_ISL_453947\|2020-0 | ···················· | ···················· | ···················· | ···················· | ···················· | ···················· | |
| C | 9611 hCoV-19/Portugal/PT0237/2020\|EPI_ISL_453951\|2020-0 | ···················· | ···················· | ···················· | ···················· | ···················· | ···················· | |
| C | 9612 hCoV-19/Portugal/PT0244/2020\|EPI_ISL_453958\|2020-0 | ···················· | ···················· | ···················· | ···················· | ···················· | ···················· | |
| C | 9613 hCoV-19/Brazil/RJ-2033/2020\|EPI_ISL_456100\|2020-04 | ···················· | ···················· | ···················· | ···················· | ···················· | ···················· | |
| C | 9614 hCoV-19/Brazil/RJ-2044/2020\|EPI_ISL_456101\|2020-04 | ···················· | ···················· | ···················· | ···················· | ···················· | ···················· | |
| C | 9615 hCoV-19/Brazil/RJ-2057/2020\|EPI_ISL_456102\|2020-04 | ···················· | ···················· | ···················· | ···················· | ···················· | ···················· | |
| C | 9616 hCoV-19/Brazil/RJ-2062/2020\|EPI_ISL_456103\|2020-04 | ···················· | ···················· | ···················· | ···················· | ···················· | ···················· | |
| C | 9617 hCoV-19/Brazil/RJ-2072/2020\|EPI_ISL_456104\|2020-04 | ···················· | ···················· | ···················· | ···················· | ···················· | ···················· | |
| C | 9618 hCoV-19/Brazil/RJ-2077/2020\|EPI_ISL_456105\|2020-04 | ···················· | ···················· | ···················· | ···················· | ···················· | ···················· | |
| C | 9619 hCoV-19/Brazil/RJ-2078/2020\|EPI_ISL_456106\|2020-04 | ···················· | ···················· | ···················· | ···················· | ···················· | ···················· | |
| C | 9620 hCoV-19/USA/NY-NYUMC834/2020\|EPI_ISL_456107\|2020-0 | ···················· | ···················· | ···················· | ···················· | ···················· | ···················· | |
| C | 9621 hCoV-19/USA/NY-NYUMC835/2020\|EPI_ISL_456108\|2020-0 | ···················· | ···················· | ···················· | ···················· | ···················· | ···················· | |
| C | 9622 hCoV-19/USA/NY-NYUMC836/2020\|EPI_ISL_456109\|2020-0 | ···················· | ···················· | ···················· | ···················· | ···················· | ···················· | |
| C | 9623 hCoV-19/USA/NY-NYUMC839/2020\|EPI_ISL_456111\|2020-0 | ···················· | ···················· | ···················· | ···················· | ···················· | ···················· | |
| C | 9624 hCoV-19/USA/NY-NYUMC840/2020\|EPI_ISL_456112\|2020-0 | ···················· | ···················· | ···················· | ···················· | ···················· | ···················· | |
| C | 9625 hCoV-19/USA/NJ-NYUMC841/2020\|EPI_ISL_456113\|2020-0 | ···················· | ···················· | ···················· | ···················· | ···················· | ···················· | |
| C | 9626 hCoV-19/New Zealand/20VR0465/2020\|EPI_ISL_456160\|2 | ···················· | ···················· | ···················· | ···················· | ···················· | ···················· | |
| C | 9627 hCoV-19/New Zealand/20VR0514/2020\|EPI_ISL_456161\|2 | ···················· | ···················· | ···················· | ···················· | ···················· | ···················· | |
| C | 9628 hCoV-19/New Zealand/20VR0712/2020\|EPI_ISL_456163\|2 | ···················· | ···················· | ···················· | ···················· | ···················· | ···················· | |
| C | 9629 hCoV-19/New Zealand/20VR0760/2020\|EPI_ISL_456165\|2 | ···················· | ···················· | ···················· | ···················· | ···················· | ···················· | |
| C | 9630 hCoV-19/New Zealand/20VR0765/2020\|EPI_ISL_456167\|2 | ···················· | ···················· | ···················· | ···················· | ···················· | ···················· | |
| C | 9631 hCoV-19/New Zealand/20VR0770/2020\|EPI_ISL_456168\|2 | ···················· | ···················· | ···················· | ···················· | ···················· | ···················· | |
| C | 9632 hCoV-19/New Zealand/20VR0901/2020\|EPI_ISL_456169\|2 | ···················· | ·············A······ | ···················· | ···················· | ···················· | ···················· | |
| C | 9633 hCoV-19/New Zealand/20VR0914/2020\|EPI_ISL_456170\|2 | ···················· | ···················· | ···················· | ···················· | ···················· | ···················· | |
| C | 9634 hCoV-19/New Zealand/20VR0933/2020\|EPI_ISL_456171\|2 | ···················· | ···················· | ···················· | ···················· | ···················· | ···················· | |
| C | 9635 hCoV-19/New Zealand/20VR0987/2020\|EPI_ISL_456172\|2 | ···················· | ···················· | ···················· | ···················· | ···················· | ···················· | |
| C | 9636 hCoV-19/New Zealand/20VR0988/2020\|EPI_ISL_456173\|2 | ···················· | ···················· | ···················· | ···················· | ···················· | ···················· | |
| C | 9637 hCoV-19/New Zealand/20VR0989/2020\|EPI_ISL_456174\|2 | ···················· | ···················· | ···················· | ···················· | ···················· | ···················· | |
| C | 9638 hCoV-19/New Zealand/20VR0990/2020\|EPI_ISL_456175\|2 | ···················· | ···················· | ···················· | ···················· | ···················· | ···················· | |
| C | 9639 hCoV-19/New Zealand/20VR0993/2020\|EPI_ISL_456176\|2 | ···················· | ···················· | ···················· | ···················· | ···················· | ···················· | |
| C | 9640 hCoV-19/New Zealand/20VR0994/2020\|EPI_ISL_456177\|2 | ···················· | ···················· | ···················· | ···················· | ···················· | ···················· | |
| C | 9641 hCoV-19/New Zealand/20VR0995/2020\|EPI_ISL_456178\|2 | ···················· | ···················· | ···················· | ···················· | ···················· | ···················· | |
| C | 9642 hCoV-19/New Zealand/20VR0996/2020\|EPI_ISL_456179\|2 | ···················· | ···················· | ···················· | ···················· | ···················· | ···················· | |
| C | 9643 hCoV-19/New Zealand/20VR0999/2020\|EPI_ISL_456180\|2 | ···················· | ···················· | ···················· | ···················· | ···················· | ···················· | |
| C | 9644 hCoV-19/New Zealand/20VR1004/2020\|EPI_ISL_456181\|2 | ···················· | ·A·················· | ···················· | ···················· | ···················· | ···················· | |
| C | 9645 hCoV-19/New Zealand/20VR1016/2020\|EPI_ISL_456182\|2 | ···················· | ···················· | ···················· | ···················· | ···················· | ···················· | |
| C | 9646 hCoV-19/New Zealand/20VR1018/2020\|EPI_ISL_456183\|2 | ···················· | ···················· | ···················· | ···················· | ···················· | ···················· | |
| C | 9647 hCoV-19/New Zealand/20VR1019/2020\|EPI_ISL_456184\|2 | ···················· | ···················· | ···················· | ···················· | ···················· | ···················· | |
| C | 9648 hCoV-19/New Zealand/20VR1020/2020\|EPI_ISL_456185\|2 | ···················· | ···················· | ···················· | ···················· | ···················· | ···················· | |
| C | 9649 hCoV-19/New Zealand/20VR1022/2020\|EPI_ISL_456186\|2 | ···················· | ···················· | ···················· | ···················· | ···················· | ···················· | |
| C | 9650 hCoV-19/New Zealand/20VR1076/2020\|EPI_ISL_456187\|2 | ···················· | ···················· | ···················· | ···················· | ···················· | ···················· | |
| C | 9651 hCoV-19/New Zealand/20VR1078/2020\|EPI_ISL_456188\|2 | ···················· | ···················· | ···················· | ···················· | ···················· | ···················· | |
| C | 9652 hCoV-19/New Zealand/20VR1109/2020\|EPI_ISL_456190\|2 | ···················· | ···················· | ···················· | ···················· | ···················· | ···················· | |
| C | 9653 hCoV-19/New Zealand/20VR1110/2020\|EPI_ISL_456191\|2 | ···················· | ···················· | ···················· | ···················· | ···················· | ···················· | |

| | | | | | | | | | |
|---|---|---|---|---|---|---|---|---|---|
| C | 9654 | hCoV-19/New Zealand/20VR1114/2020\|EPI_ISL_456192\|2 | ···················· | ···················· | ···················· | ···················· | ···················· | ···················· | |
| C | 9655 | hCoV-19/New Zealand/20VR1115/2020\|EPI_ISL_456193\|2 | ···················· | ···················· | ···················· | ···················· | ···················· | ···················· | |
| C | 9656 | hCoV-19/New Zealand/20VR1133/2020\|EPI_ISL_456194\|2 | ···················· | ···················· | ···················· | ···················· | ···················· | ···················· | |
| C | 9657 | hCoV-19/New Zealand/20VR1192/2020\|EPI_ISL_456195\|2 | ···················· | ··············C····· | ···················· | ···················· | ···················· | ···················· | |
| C | 9658 | hCoV-19/New Zealand/20VR1272/2020\|EPI_ISL_456197\|2 | ···················· | ···················· | ···················· | ···················· | ···················· | ···················· | |
| C | 9659 | hCoV-19/New Zealand/20VR1273/2020\|EPI_ISL_456198\|2 | ···················· | ···················· | ···················· | ···················· | ···················· | ···················· | |
| C | 9660 | hCoV-19/New Zealand/20VR1274/2020\|EPI_ISL_456199\|2 | ···················· | ···················· | ···················· | ···················· | ···················· | ···················· | |
| C | 9661 | hCoV-19/England/NORW-EB2F6/2020\|EPI_ISL_457416\|202 | ···················· | ···················· | ···················· | ···················· | ···················· | ···················· | |
| C | 9662 | hCoV-19/England/NORW-EB311/2020\|EPI_ISL_457417\|202 | ···················· | ···················· | ···················· | ···················· | ···················· | ···················· | |
| C | 9663 | hCoV-19/England/NORW-EB320/2020\|EPI_ISL_457418\|202 | ···················· | ···················· | ···················· | ···················· | ···················· | ···················· | |
| C | 9664 | hCoV-19/England/NORW-EB3C6/2020\|EPI_ISL_457426\|202 | ···················· | ···················· | ···················· | ···················· | ···················· | ···················· | |
| C | 9665 | hCoV-19/England/NORW-EB3D5/2020\|EPI_ISL_457427\|202 | ···················· | ···················· | ···················· | ···················· | ···················· | ···················· | |
| C | 9667 | hCoV-19/England/NORW-EB43C/2020\|EPI_ISL_457431\|202 | ···················· | ···················· | ···················· | ···················· | NNN················· | ···················· | 1 |
| C | 9668 | hCoV-19/England/NORW-EB469/2020\|EPI_ISL_457433\|202 | ···················· | ···················· | ···················· | ···················· | ···················· | ···················· | |
| C | 9669 | hCoV-19/England/NORW-EB487/2020\|EPI_ISL_457434\|202 | ···················· | ···················· | ···················· | ···················· | ···················· | ···················· | |
| C | 9670 | hCoV-19/England/NORW-EB4B4/2020\|EPI_ISL_457436\|202 | ···················· | ···················· | ···················· | ···················· | ···················· | ···················· | |
| C | 9671 | hCoV-19/England/NORW-EB4D2/2020\|EPI_ISL_457437\|202 | ···················· | ···················· | ···················· | ···················· | ···················· | ···················· | |
| C | 9672 | hCoV-19/England/NORW-EB4F0/2020\|EPI_ISL_457438\|202 | ···················· | ···················· | ···················· | ···················· | ···················· | ···················· | |
| C | 9673 | hCoV-19/England/NORW-EB52A/2020\|EPI_ISL_457440\|202 | ···················· | ···················· | ···················· | ···················· | ···················· | ···················· | |
| C | 9674 | hCoV-19/England/NORW-EB539/2020\|EPI_ISL_457441\|202 | ···················· | ···················· | ···················· | ···················· | ···················· | ···················· | |
| C | 9675 | hCoV-19/England/NORW-EB548/2020\|EPI_ISL_457442\|202 | ···················· | ···················· | ···················· | ···················· | ···················· | ···················· | |
| C | 9676 | hCoV-19/England/NORW-EB5C0/2020\|EPI_ISL_457445\|202 | ···················· | ···················· | ···················· | ···················· | ···················· | ···················· | |
| C | 9677 | hCoV-19/England/NORW-EB5DF/2020\|EPI_ISL_457446\|202 | ···················· | ···················· | ···················· | ···················· | ···················· | ···················· | |
| C | 9678 | hCoV-19/England/NORW-EB5EE/2020\|EPI_ISL_457447\|202 | ···················· | ···················· | ···················· | ···················· | ···················· | ···················· | |
| C | 9679 | hCoV-19/England/NORW-EBC07/2020\|EPI_ISL_457490\|202 | ···················· | ···················· | ···················· | ···················· | ···················· | ···················· | |
| C | 9680 | hCoV-19/England/NORW-EBC25/2020\|EPI_ISL_457491\|202 | ··G················· | ···················· | ···················· | ···················· | ···················· | ···················· | |
| C | 9681 | hCoV-19/England/NORW-EBC34/2020\|EPI_ISL_457492\|202 | ···················· | ···················· | ···················· | ···················· | ···················· | ···················· | |
| C | 9682 | hCoV-19/England/NORW-EBC43/2020\|EPI_ISL_457493\|202 | ···················· | ···················· | ···················· | ···················· | ···················· | ···················· | |
| C | 9683 | hCoV-19/England/NORW-EBC52/2020\|EPI_ISL_457494\|202 | ···················· | ···················· | ···················· | ···················· | ···················· | ···················· | |
| C | 9684 | hCoV-19/England/NORW-EBC70/2020\|EPI_ISL_457495\|202 | ···················· | ···················· | ···················· | ···················· | ···················· | ···················· | |
| C | 9685 | hCoV-19/England/NORW-EBC8F/2020\|EPI_ISL_457496\|202 | ···················· | ···················· | ···················· | ···················· | ···················· | ···················· | |
| C | 9686 | hCoV-19/England/NORW-EBC9E/2020\|EPI_ISL_457497\|202 | ···················· | ···················· | ···················· | ···················· | ···················· | ···················· | |
| C | 9687 | hCoV-19/England/NORW-EBCAD/2020\|EPI_ISL_457498\|202 | ···················· | ···················· | ···················· | ···················· | ···················· | ···················· | |
| C | 9688 | hCoV-19/England/NORW-EBCE9/2020\|EPI_ISL_457499\|202 | ···················· | ···················· | ···················· | ···················· | ···················· | ···················· | |
| C | 9689 | hCoV-19/England/BRIS-129431/2020\|EPI_ISL_458748\|20 | ···················· | ···················· | ···················· | ···················· | ···················· | ···················· | |
| C | 9690 | hCoV-19/England/BRIS-12953E/2020\|EPI_ISL_458749\|20 | ···················· | ·············A······ | ···················· | ···················· | ···················· | ···················· | |
| C | 9691 | hCoV-19/England/BRIS-128F31/2020\|EPI_ISL_458757\|20 | ···················· | ···················· | ···················· | ···················· | ···················· | ···················· | |
| C | 9692 | hCoV-19/England/BRIS-128757/2020\|EPI_ISL_458758\|20 | ···················· | ···················· | ···················· | ···················· | ···················· | ···················· | |
| C | 9693 | hCoV-19/England/BRIS-128B97/2020\|EPI_ISL_458759\|20 | ···················· | ···················· | ···················· | ···················· | ···················· | ···················· | |
| C | 9694 | hCoV-19/England/BRIS-128E70/2020\|EPI_ISL_458760\|20 | ···················· | ···················· | ···················· | ···················· | ···················· | ···················· | |
| C | 9695 | hCoV-19/England/BRIS-1291EF/2020\|EPI_ISL_458761\|20 | ···················· | ···················· | ···················· | ···················· | ···················· | ···················· | |
| C | 9696 | hCoV-19/England/BRIS-12963B/2020\|EPI_ISL_458762\|20 | ···················· | ···················· | ···················· | ···················· | ···················· | ···················· | |
| C | 9697 | hCoV-19/England/BRIS-128FE6/2020\|EPI_ISL_458763\|20 | ···················· | ···················· | ···················· | ···················· | ···················· | ···················· | |
| C | 9698 | hCoV-19/England/BRIS-1295F2/2020\|EPI_ISL_458764\|20 | ···················· | ···················· | ···················· | ···················· | ···················· | ···················· | |
| C | 9699 | hCoV-19/England/BRIS-1295A7/2020\|EPI_ISL_458765\|20 | ···················· | ···················· | ···················· | ···················· | ···················· | ···················· | |
| C | 9700 | hCoV-19/England/BRIS-1294E6/2020\|EPI_ISL_458766\|20 | ···················· | ···················· | ···················· | ···················· | ···················· | ···················· | |
| C | 9701 | hCoV-19/England/BRIS-12946E/2020\|EPI_ISL_458767\|20 | ···················· | ···················· | ···················· | ···················· | ···················· | ···················· | |
| C | 9702 | hCoV-19/England/BRIS-129817/2020\|EPI_ISL_458768\|20 | ···················· | ···················· | ···················· | ···················· | ···················· | ···················· | |
| C | 9703 | hCoV-19/England/BRIS-1294AA/2020\|EPI_ISL_458769\|20 | ···················· | ···················· | ···················· | ···················· | ···················· | ···················· | |
| C | 9704 | hCoV-19/England/BRIS-1296D1/2020\|EPI_ISL_458770\|20 | ···················· | ···················· | ···················· | ···················· | ···················· | ···················· | |
| C | 9705 | hCoV-19/England/BRIS-128E25/2020\|EPI_ISL_458771\|20 | ···················· | ···················· | ···················· | ···················· | ···················· | ···················· | |
| C | 9706 | hCoV-19/England/BRIS-128A30/2020\|EPI_ISL_458772\|20 | ···················· | ···················· | ···················· | ···················· | ···················· | ···················· | |
| C | 9707 | hCoV-19/England/BRIS-128933/2020\|EPI_ISL_458773\|20 | ···················· | ···················· | ···················· | ···················· | ···················· | ···················· | |
| C | 9708 | hCoV-19/England/BRIS-128BA6/2020\|EPI_ISL_458774\|20 | ···················· | ···················· | ···················· | ···················· | ···················· | ···················· | |
| C | 9709 | hCoV-19/England/BRIS-1291B2/2020\|EPI_ISL_458775\|20 | ···················· | ···················· | ···················· | ···················· | ···················· | ···················· | |
| C | 9710 | hCoV-19/England/BRIS-128D64/2020\|EPI_ISL_458777\|20 | ···················· | ····A··············· | ···················· | ···················· | ···················· | ···················· | |
| C | 9711 | hCoV-19/England/BRIS-129695/2020\|EPI_ISL_458778\|20 | ···················· | ···················· | ···················· | ···················· | ···················· | ···················· | |
| C | 9712 | hCoV-19/England/SHEF-C0EF3/2020\|EPI_ISL_462000\|202 | ···················· | ···················· | ···················· | ···················· | ···················· | ···················· | |
| C | 9713 | hCoV-19/England/SHEF-C1108/2020\|EPI_ISL_462001\|202 | ···················· | ···················· | ···················· | ···················· | ···················· | ···················· | |
| C | 9714 | hCoV-19/England/SHEF-C15A2/2020\|EPI_ISL_462002\|202 | ···················· | ···················· | ···················· | ···················· | ···················· | ···················· | |
| C | 9715 | hCoV-19/England/SHEF-C1ED4/2020\|EPI_ISL_462003\|202 | ···················· | ···················· | ···················· | ···················· | ···················· | ···················· | |
| C | 9716 | hCoV-19/England/SHEF-C1F59/2020\|EPI_ISL_462004\|202 | ···················· | ···················· | ···················· | ···················· | ···················· | ···················· | |
| C | 9717 | hCoV-19/England/SHEF-C2459/2020\|EPI_ISL_462005\|202 | ···················· | ···················· | ···················· | ···················· | ···················· | ···················· | |
| C | 9718 | hCoV-19/England/SHEF-C2D30/2020\|EPI_ISL_462007\|202 | ···················· | ···················· | ···················· | ···················· | ···················· | ···················· | |
| C | 9719 | hCoV-19/England/SHEF-C2DE5/2020\|EPI_ISL_462008\|202 | ···················· | ···················· | ···················· | ···················· | ···················· | ···················· | |
| C | 9720 | hCoV-19/England/SHEF-C31CA/2020\|EPI_ISL_462009\|202 | ···················· | ···················· | ···················· | ···················· | ···················· | ···················· | |
| C | 9721 | hCoV-19/England/SHEF-C3B17/2020\|EPI_ISL_462010\|202 | ···················· | ···················· | ···················· | ···················· | ···················· | ···················· | |
| C | 9722 | hCoV-19/England/SHEF-C3B71/2020\|EPI_ISL_462011\|202 | ···················· | ···················· | ···················· | ···················· | ···················· | ···················· | |
| C | 9723 | hCoV-19/England/SHEF-C4F56/2020\|EPI_ISL_462016\|202 | ···················· | ···················· | ···················· | ···················· | ···················· | ···················· | |
| C | 9724 | hCoV-19/England/SHEF-C50BC/2020\|EPI_ISL_462017\|202 | ···················· | ···················· | ···················· | ···················· | ···················· | ···················· | |
| C | 9725 | hCoV-19/England/SHEF-C54FC/2020\|EPI_ISL_462018\|202 | ···················· | ···················· | ···················· | ···················· | ···················· | ···················· | |
| C | 9726 | hCoV-19/England/SHEF-C590C/2020\|EPI_ISL_462020\|202 | ···················· | ···················· | ···················· | ···················· | ···················· | ···················· | |
| C | 9727 | hCoV-19/England/SHEF-C5F64/2020\|EPI_ISL_462021\|202 | ···················· | ···················· | ···················· | ···················· | ···················· | ···················· | |
| C | 9728 | hCoV-19/England/SHEF-C6376/2020\|EPI_ISL_462023\|202 | ···················· | ···················· | ···················· | ···················· | ···················· | ···················· | |
| C | 9733 | hCoV-19/Colombia/QUI-INS-79911/2020\|EPI_ISL_456127 | ···················· | ···················· | ···················· | ···················· | ···················· | ···················· | |
| C | 9734 | hCoV-19/Colombia/VAC-INS-79943/2020\|EPI_ISL_456138 | ···················· | ···················· | ···················· | ···················· | ···················· | ···················· | |
| C | 9735 | hCoV-19/Colombia/AMA-INS-103039/2020\|EPI_ISL_45614 | ···················· | ···················· | ···················· | ···················· | ···················· | ···················· | |
| C | 9736 | hCoV-19/Colombia/ATL-INS-79743/2020\|EPI_ISL_456145 | ···················· | ···················· | ···················· | ···················· | ···················· | ···················· | |

| | | | | | | | | |
|---|---|---|---|---|---|---|---|---|
| C | 9737 hCoV-19/Colombia/ANT-INS-82457/2020\|EPI_ISL_456146 | .................... | .................... | .................... | .................... | .................... | .................... | |
| C | 9738 hCoV-19/Colombia/VAC-INS-96183/2020\|EPI_ISL_456147 | .................... | .................... | .................... | .................... | .................... | .................... | |
| C | 9739 hCoV-19/Colombia/BOL-INS-79890/2020\|EPI_ISL_456148 | .................... | .................... | .................... | .................... | .................... | .................... | |
| C | 9740 hCoV-19/Colombia/SAN-INS-80123/2020\|EPI_ISL_456149 | .................... | .................... | .................... | .................... | .................... | .................... | |
| C | 9741 hCoV-19/Colombia/VAC-INS-79965/2020\|EPI_ISL_456150 | .................... | .................... | .................... | .................... | .................... | .................... | |
| C | 9742 hCoV-19/Colombia/VAC-INS-82320/2020\|EPI_ISL_456151 | .................... | .................... | .................... | .................... | .................... | .................... | |
| C | 9743 hCoV-19/Colombia/VAC-INS-96176/2020\|EPI_ISL_456152 | .................... | .................... | .................... | .................... | .................... | .................... | |
| C | 9744 hCoV-19/Colombia/MAG-INS-101935/2020\|EPI_ISL_45615 | .................... | .................... | .................... | .................... | .................... | .................... | |
| C | 9745 hCoV-19/Colombia/NAR-INS-103303/2020\|EPI_ISL_45615 | .................... | .................... | .................... | .................... | .................... | .................... | |
| C | 9746 hCoV-19/Colombia/RIS-INS-80245/2020\|EPI_ISL_456155 | .................... | .................... | .................... | .................... | .................... | .................... | |
| C | 9747 hCoV-19/New Zealand/20VR0462/2020\|EPI_ISL_456159\|2 | .................... | .................... | .................... | .................... | .................... | .................... | |
| C | 9748 hCoV-19/England/NORW-EB00B/2020\|EPI_ISL_457405\|202 | .................... | .................... | .................... | .................... | .................... | .................... | |
| C | 9749 hCoV-19/England/NORW-EB01A/2020\|EPI_ISL_457406\|202 | .................... | .................... | .................... | .................... | .................... | .................... | |
| C | 9750 hCoV-19/England/NORW-EB029/2020\|EPI_ISL_457407\|202 | .................... | .................... | .................... | .................... | .................... | .................... | |
| C | 9751 hCoV-19/England/NORW-EB038/2020\|EPI_ISL_457408\|202 | .................... | .................... | .................... | .................... | .................... | .................... | |
| C | 9752 hCoV-19/England/NORW-EB047/2020\|EPI_ISL_457409\|202 | .................... | .................... | .................... | .................... | .................... | .................... | |
| C | 9753 hCoV-19/England/NORW-EB232/2020\|EPI_ISL_457411\|202 | .................... | .................... | .................... | .................... | .................... | .................... | |
| C | 9754 hCoV-19/England/NORW-EB26F/2020\|EPI_ISL_457412\|202 | .................... | .................... | .................... | .................... | .................... | .................... | |
| C | 9755 hCoV-19/England/NORW-EB28D/2020\|EPI_ISL_457413\|202 | .................... | .................... | .................... | .................... | .................... | .................... | |
| C | 9756 hCoV-19/England/NORW-EB29C/2020\|EPI_ISL_457414\|202 | .................... | .................... | .................... | .................... | .................... | .................... | |
| C | 9757 hCoV-19/England/NORW-EB2AB/2020\|EPI_ISL_457415\|202 | .................... | .................... | .................... | .................... | .................... | .................... | |
| C | 9758 hCoV-19/England/NORW-EB33F/2020\|EPI_ISL_457419\|202 | .................... | .................... | .................... | .................... | .................... | .................... | |
| C | 9759 hCoV-19/England/NORW-EB34E/2020\|EPI_ISL_457420\|202 | .................... | .................... | .................... | .................... | .................... | .................... | |
| C | 9760 hCoV-19/England/NORW-EB37B/2020\|EPI_ISL_457423\|202 | .................... | .................... | .................... | .................... | .................... | .................... | |
| C | 9761 hCoV-19/England/NORW-EB399/2020\|EPI_ISL_457425\|202 | .................... | .................... | .................... | .................... | .................... | .................... | |
| C | 9762 hCoV-19/England/NORW-EB609/2020\|EPI_ISL_457448\|202 | .................... | .................... | .................... | .................... | .................... | .................... | |
| C | 9763 hCoV-19/England/NORW-EB618/2020\|EPI_ISL_457449\|202 | .................... | .................... | .................... | .................... | .................... | .................... | |
| C | 9764 hCoV-19/England/NORW-EB636/2020\|EPI_ISL_457451\|202 | .................... | .................... | .................... | .................... | .................... | .................... | |
| C | 9766 hCoV-19/England/NORW-EB900/2020\|EPI_ISL_457455\|202 | .................... | .................... | .................... | .................... | .................... | .................... | |
| C | 9767 hCoV-19/England/NORW-EB91F/2020\|EPI_ISL_457456\|202 | .................... | .................... | .................... | .................... | .................... | .................... | |
| C | 9768 hCoV-19/England/NORW-EB93D/2020\|EPI_ISL_457457\|202 | .................... | .................... | .................... | .................... | .................... | .................... | |
| C | 9769 hCoV-19/England/NORW-EB94C/2020\|EPI_ISL_457458\|202 | .................... | .................... | .................... | .................... | .................... | .................... | |
| C | 9770 hCoV-19/England/NORW-EB95B/2020\|EPI_ISL_457459\|202 | .................... | .................... | .................... | .................... | .................... | .................... | |
| C | 9771 hCoV-19/England/NORW-EB979/2020\|EPI_ISL_457460\|202 | .................... | .................... | .................... | .................... | .................... | .................... | |
| C | 9772 hCoV-19/England/NORW-EB997/2020\|EPI_ISL_457461\|202 | .................... | ...................A | .................... | .................... | .................... | .................... | 1 |
| C | 9773 hCoV-19/England/NORW-EB9A6/2020\|EPI_ISL_457462\|202 | .................... | .................... | .................... | .................... | .................... | .................... | |
| C | 9774 hCoV-19/England/NORW-EB9B5/2020\|EPI_ISL_457463\|202 | .................... | .................... | .................... | .................... | .................... | .................... | |
| C | 9775 hCoV-19/England/NORW-EB9C4/2020\|EPI_ISL_457464\|202 | .................... | .................... | .................... | .................... | .................... | .................... | |
| C | 9776 hCoV-19/England/NORW-EB9D3/2020\|EPI_ISL_457465\|202 | .................... | .................... | .................... | .................... | .................... | .................... | |
| C | 9777 hCoV-19/England/NORW-EBA3A/2020\|EPI_ISL_457467\|202 | .................... | ...................A | .................... | .................... | .................... | .................... | 1 |
| C | 9778 hCoV-19/England/NORW-EBA49/2020\|EPI_ISL_457468\|202 | .................... | .................... | .................... | .................... | .................... | .................... | |
| C | 9779 hCoV-19/England/NORW-EBA67/2020\|EPI_ISL_457469\|202 | .................... | .................... | .................... | .................... | .................... | .................... | |
| C | 9780 hCoV-19/England/NORW-EBA76/2020\|EPI_ISL_457470\|202 | .................... | .................... | .................... | .................... | .................... | .................... | |
| C | 9781 hCoV-19/England/NORW-EBA85/2020\|EPI_ISL_457471\|202 | .................... | .................... | .................... | .................... | .................... | .................... | |
| C | 9782 hCoV-19/England/NORW-EBA94/2020\|EPI_ISL_457472\|202 | .................... | .................... | .................... | .................... | .................... | .................... | |
| C | 9783 hCoV-19/England/NORW-EBAA3/2020\|EPI_ISL_457473\|202 | .................... | .................... | .................... | .................... | .................... | .................... | |
| C | 9784 hCoV-19/England/NORW-EBAC1/2020\|EPI_ISL_457474\|202 | .................... | .................... | .................... | .................... | .................... | .................... | |
| C | 9785 hCoV-19/England/NORW-EBAD0/2020\|EPI_ISL_457475\|202 | .................... | .................... | .................... | .................... | .................... | .................... | |
| C | 9786 hCoV-19/England/NORW-EBAEF/2020\|EPI_ISL_457476\|202 | .................... | .................... | .................... | .................... | .................... | .................... | |
| C | 9787 hCoV-19/England/NORW-EBAFE/2020\|EPI_ISL_457477\|202 | .................... | .................... | .................... | .................... | .................... | .................... | |
| C | 9788 hCoV-19/England/NORW-EBB28/2020\|EPI_ISL_457478\|202 | .................... | .................... | .................... | .................... | .................... | .................... | |
| C | 9789 hCoV-19/England/NORW-EBB37/2020\|EPI_ISL_457479\|202 | .................... | .................... | .................... | .................... | .................... | .................... | |
| C | 9790 hCoV-19/England/NORW-EBB46/2020\|EPI_ISL_457480\|202 | .................... | .................... | .................... | .................... | .................... | .................... | |
| C | 9791 hCoV-19/England/NORW-EBB64/2020\|EPI_ISL_457482\|202 | .................... | .................... | .................... | .................... | .................... | .................... | |
| C | 9792 hCoV-19/England/NORW-EBB73/2020\|EPI_ISL_457483\|202 | .................... | .................... | .................... | .................... | .................... | .................... | |
| C | 9793 hCoV-19/England/NORW-EBB82/2020\|EPI_ISL_457484\|202 | .................... | .................... | .................... | .................... | .................... | .................... | |
| C | 9794 hCoV-19/England/NORW-EBBA0/2020\|EPI_ISL_457485\|202 | ..................T. | .................... | .................... | .................... | .................... | .................... | 1 |
| C | 9795 hCoV-19/England/NORW-EBBBF/2020\|EPI_ISL_457486\|202 | .................... | .................... | .................... | .................... | .................... | .................... | |
| C | 9796 hCoV-19/England/NORW-EBBCE/2020\|EPI_ISL_457487\|202 | .................... | .................... | .................... | .................... | .................... | .................... | |
| C | 9797 hCoV-19/England/NORW-EBBEC/2020\|EPI_ISL_457488\|202 | .................... | .................... | .................... | .................... | .................... | .................... | |
| C | 9798 hCoV-19/England/NORW-EBBFB/2020\|EPI_ISL_457489\|202 | .................... | .................... | .................... | .................... | .................... | .................... | |
| C | 9799 hCoV-19/England/BRIS-128D73/2020\|EPI_ISL_458736\|20 | .................... | ...A................ | .................... | .................... | .................... | .................... | |
| C | 9800 hCoV-19/England/BRIS-128854/2020\|EPI_ISL_458739\|20 | .................... | .................... | .................... | .................... | .................... | .................... | |
| C | 9801 hCoV-19/England/BRIS-129246/2020\|EPI_ISL_458743\|20 | .................... | .................... | .................... | .................... | .................... | .................... | |
| C | 9802 hCoV-19/England/BRIS-128B3D/2020\|EPI_ISL_458745\|20 | .................... | .................... | .................... | .................... | .................... | .................... | |
| C | 9803 hCoV-19/England/BRIS-128C58/2020\|EPI_ISL_458746\|20 | .................... | .................... | .................... | .................... | .................... | .................... | |
| C | 9804 hCoV-19/England/BRIS-128EF8/2020\|EPI_ISL_458750\|20 | .................... | .................... | .................... | .................... | .................... | .................... | |
| C | 9805 hCoV-19/England/BRIS-128CB2/2020\|EPI_ISL_458751\|20 | .................... | .................... | .................... | .................... | .................... | .................... | |
| C | 9806 hCoV-19/England/BRIS-128818/2020\|EPI_ISL_458752\|20 | .................... | .................... | .................... | .................... | .................... | .................... | |
| C | 9807 hCoV-19/England/BRIS-1287FD/2020\|EPI_ISL_458755\|20 | .................... | .................... | .................... | .................... | .................... | .................... | |
| C | 9808 hCoV-19/England/BRIS-129237/2020\|EPI_ISL_458756\|20 | .................... | .................... | .................... | .................... | .................... | .................... | |
| C | 9809 hCoV-19/England/BRIS-128AA9/2020\|EPI_ISL_458779\|20 | .................... | .............A...... | .................... | .................... | .................... | .................... | |
| C | 9810 hCoV-19/England/BRIS-128EE9/2020\|EPI_ISL_458781\|20 | .................... | .................... | .................... | .................... | .................... | .................... | |
| C | 9811 hCoV-19/England/BRIS-129167/2020\|EPI_ISL_458782\|20 | .................... | .................... | .................... | .................... | .................... | .................... | |
| C | 9812 hCoV-19/England/BRIS-12913A/2020\|EPI_ISL_458783\|20 | .................... | .................... | .................... | .................... | .................... | .................... | |
| C | 9813 hCoV-19/England/BRIS-1292DD/2020\|EPI_ISL_458784\|20 | .................... | .................... | .................... | .................... | .................... | .................... | |
| C | 9814 hCoV-19/England/BRIS-1292EC/2020\|EPI_ISL_458785\|20 | .................... | .................... | .................... | .................... | .................... | .................... | |
| C | 9815 hCoV-19/England/BRIS-129334/2020\|EPI_ISL_458787\|20 | .................... | .................... | .................... | .................... | .................... | .................... | |

| | | | | | | | | | |
|---|---|---|---|---|---|---|---|---|---|
| C | 9816 | hCoV-19/England/BRIS-1292A0/2020\|EPI_ISL_458791\|20 | ···················· | ···················· | ···················· | ···················· | ···················· | ···················· | |
| C | 9817 | hCoV-19/England/BRIS-12912B/2020\|EPI_ISL_458792\|20 | ···················· | ···················· | ···················· | ···················· | ···················· | ···················· | |
| C | 9818 | hCoV-19/England/BRIS-12870C/2020\|EPI_ISL_458793\|20 | ···················· | ···················· | ···················· | ···················· | ···················· | ···················· | |
| C | 9819 | hCoV-19/England/BRIS-12954D/2020\|EPI_ISL_458794\|20 | ···················· | ·············A······ | ···················· | ···················· | ···················· | ···················· | |
| C | 9820 | hCoV-19/England/BRIS-128C94/2020\|EPI_ISL_458796\|20 | ···················· | ···················· | ···················· | ···················· | ···················· | ···················· | |
| C | 9821 | hCoV-19/England/BRIS-128ECB/2020\|EPI_ISL_458797\|20 | ···················· | ···················· | ···················· | ···················· | ···················· | ···················· | |
| C | 9822 | hCoV-19/England/BRIS-128A9A/2020\|EPI_ISL_458798\|20 | ···················· | ···················· | ···················· | ···················· | ···················· | ···················· | |
| C | 9823 | hCoV-19/England/BRIS-128863/2020\|EPI_ISL_458799\|20 | ···················· | ···················· | ···················· | ···················· | ···················· | ···················· | |
| D | 1 | hCoV-19/Shaoxing/01/2020\|EPI_ISL_463889\|2020-01-27 | ···················· | ···················· | ···················· | ···················· | ···················· | ···················· | |
| D | 2 | hCoV-19/Shaoxing/02/2020\|EPI_ISL_463894\|2020-01-30 | ···················· | ···················· | ···················· | ···················· | ···················· | ···················· | |
| D | 3 | hCoV-19/Shaoxing/03/2020\|EPI_ISL_463895\|2020-01-30 | ···················· | ···················· | ···················· | ···················· | ···················· | ···················· | |
| D | 4 | hCoV-19/Shaoxing/04/2020\|EPI_ISL_463896\|2020-01-31 | ···················· | ···················· | ···················· | ···················· | ···················· | ···················· | |
| D | 5 | hCoV-19/Shaoxing/05/2020\|EPI_ISL_463897\|2020-01-31 | ···················· | ···················· | ···················· | ···················· | ···················· | ···················· | |
| D | 6 | hCoV-19/England/20149001104/2020\|EPI_ISL_466081\|20 | ···················· | ···················· | ···················· | ···················· | ···N···N···N········ | ···················· | 1 |
| D | 8 | hCoV-19/England/20149001804/2020\|EPI_ISL_466084\|20 | ···················· | ···················· | ···················· | ···················· | ···················· | ···················· | |
| D | 9 | hCoV-19/England/20149002404/2020\|EPI_ISL_466086\|20 | ···················· | ···················· | ···················· | ···················· | ···················· | ···················· | |
| D | 10 | hCoV-19/England/20149002704/2020\|EPI_ISL_466088\|20 | ···················· | ···················· | ···················· | ···················· | ···················· | ···················· | |
| D | 11 | hCoV-19/England/20149003004/2020\|EPI_ISL_466089\|20 | ···················· | ···················· | ···················· | ···················· | ···················· | ···················· | |
| D | 12 | hCoV-19/England/20149003104/2020\|EPI_ISL_466090\|20 | ···················· | ···················· | ···················· | ···················· | ···················· | ···················· | |
| D | 13 | hCoV-19/England/20149003304/2020\|EPI_ISL_466092\|20 | ···················· | ···················· | ···················· | ···················· | ···················· | ···················· | |
| D | 14 | hCoV-19/England/20149003604/2020\|EPI_ISL_466094\|20 | ···················· | ···················· | ···················· | ···················· | ···················· | ···················· | |
| D | 18 | hCoV-19/England/20149004004/2020\|EPI_ISL_466098\|20 | ···················· | ···················· | ···················· | ···················· | ···················· | ···················· | |
| D | 19 | hCoV-19/England/20149004104/2020\|EPI_ISL_466099\|20 | ···················· | ···················· | ···················· | ···················· | ···················· | ···················· | |
| D | 22 | hCoV-19/England/20149004804/2020\|EPI_ISL_466103\|20 | ···················· | ···················· | ···················· | ···················· | ···················· | ···················· | |
| D | 23 | hCoV-19/England/20149005204/2020\|EPI_ISL_466107\|20 | ···················· | ···················· | ···················· | ···················· | ···················· | ···················· | |
| D | 24 | hCoV-19/England/20149005404/2020\|EPI_ISL_466109\|20 | ···················· | ···················· | ···················· | ···················· | ·NN················· | ···················· | |
| D | 25 | hCoV-19/England/20149005504/2020\|EPI_ISL_466110\|20 | ···················· | ···················· | ···················· | ···················· | ···················· | ··············N····· | |
| D | 26 | hCoV-19/England/20149005904/2020\|EPI_ISL_466113\|20 | ···················· | ···················· | ···················· | ···················· | ···················· | ···················· | |
| D | 27 | hCoV-19/England/20149006104/2020\|EPI_ISL_466115\|20 | ···················· | ···················· | ···················· | ···················· | ···················· | ···················· | |
| D | 28 | hCoV-19/USA/NY-NYUMC907/2020\|EPI_ISL_467400\|2020-0 | ···················· | ···················· | ···················· | ···················· | ···················· | ···················· | |
| D | 29 | hCoV-19/USA/NY-NYUMC912/2020\|EPI_ISL_467405\|2020-0 | ···················· | ···················· | ···················· | ···················· | ···················· | ···················· | |
| D | 30 | hCoV-19/USA/NY-NYUMC916/2020\|EPI_ISL_467409\|2020-0 | ···················· | ···················· | ···················· | ···················· | ···················· | ···················· | |
| D | 31 | hCoV-19/USA/NY-NYUMC918/2020\|EPI_ISL_467411\|2020-0 | ···················· | ···················· | ···················· | ···················· | ···················· | ···················· | |
| D | 32 | hCoV-19/USA/NY-NYUMC919/2020\|EPI_ISL_467412\|2020-0 | ···················· | ···················· | ···················· | ···················· | ···················· | ···················· | |
| D | 33 | hCoV-19/USA/NY-NYUMC920/2020\|EPI_ISL_467413\|2020-0 | ···················· | ···················· | ···················· | ···················· | ···················· | ···················· | |
| D | 34 | hCoV-19/USA/NY-NYUMC921/2020\|EPI_ISL_467414\|2020-0 | ···················· | ···················· | ···················· | ···················· | ···················· | ···················· | |
| D | 35 | hCoV-19/USA/NY-NYUMC923/2020\|EPI_ISL_467416\|2020-0 | ···················· | ···················· | ···················· | ···················· | ···················· | ···················· | |
| D | 36 | hCoV-19/USA/NY-NYUMC924/2020\|EPI_ISL_467417\|2020-0 | ···················· | ···················· | ···················· | ···················· | ···················· | ···················· | |
| D | 37 | hCoV-19/USA/NY-NYUMC925/2020\|EPI_ISL_467418\|2020-0 | ···················· | ·A·················· | ···················· | ···················· | ···················· | ···················· | |
| D | 38 | hCoV-19/USA/NY-NYUMC927/2020\|EPI_ISL_467420\|2020-0 | ···················· | ···················· | ···················· | ···················· | ···················· | ···················· | |
| D | 39 | hCoV-19/USA/NY-NYUMC928/2020\|EPI_ISL_467421\|2020-0 | ···················· | ···················· | ···················· | ···················· | ···················· | ···················· | |
| D | 40 | hCoV-19/USA/NY-NYUMC929/2020\|EPI_ISL_467422\|2020-0 | ···················· | ···················· | ···················· | ···················· | ···················· | ···················· | |
| D | 41 | hCoV-19/Canada/BC_13704688/2020\|EPI_ISL_467423\|202 | ···················· | ···················· | ···················· | ···················· | ···················· | ···················· | |
| D | 42 | hCoV-19/Canada/BC_13705728/2020\|EPI_ISL_467424\|202 | ···················· | ···················· | ···················· | ···················· | ···················· | ···················· | |
| D | 43 | hCoV-19/Canada/BC_13949840/2020\|EPI_ISL_467425\|202 | ···················· | ···················· | ···················· | ···················· | ···················· | ···················· | |
| D | 44 | hCoV-19/Canada/BC_14260476/2020\|EPI_ISL_467426\|202 | ···················· | ···················· | ···················· | ···················· | ···················· | ···················· | |
| D | 45 | hCoV-19/Canada/BC_14282914/2020\|EPI_ISL_467427\|202 | ···················· | ···················· | ···················· | ···················· | ···················· | ···················· | |
| D | 46 | hCoV-19/Canada/BC_14592296/2020\|EPI_ISL_467428\|202 | ···················· | ···················· | ···················· | ···················· | ···················· | ···················· | |
| D | 47 | hCoV-19/Canada/BC_14592943/2020\|EPI_ISL_467429\|202 | ···················· | ···················· | ···················· | ···················· | ···················· | ···················· | |
| D | 48 | hCoV-19/Harbin/HRB-25/2020\|EPI_ISL_467430\|2020-03- | ···················· | ···················· | ···················· | ···················· | ···················· | ···················· | |
| D | 49 | hCoV-19/South Africa/KRISP-0001/2020\|EPI_ISL_46743 | ···················· | ···················· | ···················· | ···················· | ···················· | ···················· | |
| D | 50 | hCoV-19/South Africa/KRISP-0016/2020\|EPI_ISL_46743 | ···················· | ···················· | ···················· | ···················· | ···················· | ···················· | |
| D | 51 | hCoV-19/South Africa/KRISP-0017/2020\|EPI_ISL_46743 | ···················· | ···················· | ···················· | ···················· | ···················· | ···················· | |
| D | 52 | hCoV-19/South Africa/KRISP-0019/2020\|EPI_ISL_46743 | ···················· | ···················· | ···················· | ···················· | ···················· | ···················· | |
| D | 53 | hCoV-19/South Africa/KRISP-0021/2020\|EPI_ISL_46743 | ···················· | ···················· | ···················· | ···················· | ···················· | ···················· | |
| D | 54 | hCoV-19/South Africa/KRISP-0026/2020\|EPI_ISL_46743 | ···················· | ···················· | ···················· | ···················· | ···················· | ···················· | |
| D | 55 | hCoV-19/South Africa/KRISP-0035/2020\|EPI_ISL_46744 | ···················· | ···················· | ···················· | ···················· | ···················· | ···················· | |
| D | 56 | hCoV-19/South Africa/KRISP-0040/2020\|EPI_ISL_46744 | ···················· | ···················· | ···················· | ···················· | ···················· | ···················· | |
| D | 57 | hCoV-19/South Africa/KRISP-0041/2020\|EPI_ISL_46744 | ···················· | ···················· | ···················· | ···················· | ···················· | ···················· | |
| D | 58 | hCoV-19/South Africa/KRISP-0055/2020\|EPI_ISL_46744 | ···················· | ···················· | ···················· | ···················· | ···················· | ···················· | |
| D | 59 | hCoV-19/South Africa/KRISP-0056/2020\|EPI_ISL_46744 | ···················· | ···················· | ···················· | ···················· | ···················· | ···················· | |
| D | 60 | hCoV-19/South Africa/KRISP-0057/2020\|EPI_ISL_46744 | ···················· | ···················· | ···················· | ···················· | ···················· | ···················· | |
| D | 61 | hCoV-19/Canada/BC_18143554/2020\|EPI_ISL_468700\|202 | ···················· | ···················· | ···················· | ···················· | ···················· | ···················· | |
| D | 62 | hCoV-19/USA/LA-BIE-001/2020\|EPI_ISL_468701\|2020-04 | ···················· | ···················· | ···················· | ···················· | ···················· | ···················· | |
| D | 63 | hCoV-19/USA/LA-BIE-002/2020\|EPI_ISL_468702\|2020-04 | ···················· | ···················· | ···················· | ···················· | ···················· | ···················· | |
| D | 64 | hCoV-19/USA/LA-BIE-003/2020\|EPI_ISL_468703\|2020-04 | ···················· | ···················· | ···················· | ···················· | ···················· | ···················· | |
| D | 65 | hCoV-19/USA/LA-BIE-009/2020\|EPI_ISL_468705\|2020-04 | ···················· | ···················· | ···················· | ···················· | ···················· | ···················· | |
| D | 66 | hCoV-19/USA/LA-BIE-011/2020\|EPI_ISL_468706\|2020-04 | ···················· | ···················· | ···················· | ···················· | ···················· | ···················· | |
| D | 67 | hCoV-19/USA/LA-BIE-015/2020\|EPI_ISL_468707\|2020-04 | ···················· | ···················· | ···················· | ···················· | ···················· | ···················· | |
| D | 68 | hCoV-19/USA/LA-BIE-016/2020\|EPI_ISL_468708\|2020-04 | ···················· | ···················· | ···················· | ···················· | ···················· | ···················· | |
| D | 69 | hCoV-19/USA/LA-BIE-018/2020\|EPI_ISL_468709\|2020-04 | ···················· | ···················· | ···················· | ···················· | ···················· | ···················· | |
| D | 70 | hCoV-19/USA/LA-BIE-019/2020\|EPI_ISL_468710\|2020-04 | ···················· | ···················· | ···················· | ···················· | ···················· | ···················· | |
| D | 71 | hCoV-19/USA/LA-BIE-020/2020\|EPI_ISL_468711\|2020-04 | ···················· | ···················· | ···················· | ···················· | ···················· | ···················· | |
| D | 72 | hCoV-19/USA/LA-BIE-022/2020\|EPI_ISL_468712\|2020-04 | ···················· | ···················· | ···················· | ···················· | ···················· | ···················· | |
| D | 73 | hCoV-19/USA/LA-BIE-023/2020\|EPI_ISL_468713\|2020-04 | ···················· | ···················· | ···················· | ···················· | ···················· | ···················· | |
| D | 74 | hCoV-19/USA/LA-BIE-025/2020\|EPI_ISL_468714\|2020-04 | ···················· | ···················· | ···················· | ···················· | ···················· | ···················· | |
| D | 75 | hCoV-19/USA/LA-BIE-026/2020\|EPI_ISL_468715\|2020-04 | ···················· | ···················· | ···················· | ···················· | ···················· | ···················· | |
| D | 76 | hCoV-19/USA/LA-BIE-027/2020\|EPI_ISL_468716\|2020-04 | ···················· | ···················· | ···················· | ···················· | ···················· | ···················· | |

| | | | | | | | | |
|---|---|---|---|---|---|---|---|---|
| D | 77 hCoV-19/USA/LA-BIE-028/2020\|EPI_ISL_468717\|2020-04 | ···················· | ···················· | ···················· | ···················· | ···················· | ···················· | |
| D | 78 hCoV-19/USA/FL-UF-13/2020\|EPI_ISL_468718\|2020-05-0 | ···················· | ···················· | ···················· | ···················· | ···················· | ···················· | |
| D | 79 hCoV-19/USA/FL-UF-14/2020\|EPI_ISL_468719\|2020-04-2 | ···················· | ···················· | ···················· | ···················· | ···················· | ···················· | |
| D | 80 hCoV-19/USA/FL-UF-17/2020\|EPI_ISL_468720\|2020-04-2 | ···················· | ···················· | ···················· | ···················· | ···················· | ···················· | |
| D | 81 hCoV-19/USA/FL-UF-15/2020\|EPI_ISL_468721\|2020-04-1 | ···················· | ···················· | ···················· | ···················· | ···················· | ···················· | |
| D | 82 hCoV-19/USA/FL-UF-18/2020\|EPI_ISL_468722\|2020-04-0 | ···················· | ···················· | ···················· | ···················· | ···················· | ···················· | |
| D | 83 hCoV-19/USA/FL-UF-16/2020\|EPI_ISL_468723\|2020-04-2 | ···················· | ···················· | ···················· | ···················· | ···················· | ···················· | |
| D | 84 hCoV-19/Shanghai/CH-03/2020\|EPI_ISL_468726\|2020-01 | ···················· | ···················· | ···················· | ···················· | ···················· | ···················· | |
| D | 85 hCoV-19/Belgium/UGent-76/2020\|EPI_ISL_468727\|2020- | ···················· | ···················· | ···················· | ···················· | ···················· | ···················· | |
| D | 86 hCoV-19/Belgium/UGent-77/2020\|EPI_ISL_468728\|2020- | ···················· | ···················· | ···················· | ···················· | ···················· | ···················· | |
| D | 89 hCoV-19/Belgium/UGent-82/2020\|EPI_ISL_468731\|2020- | ···················· | ···················· | ···················· | ···················· | ···················· | ···················· | |
| D | 90 hCoV-19/Belgium/UGent-83/2020\|EPI_ISL_468732\|2020- | ···················· | ···················· | ···················· | ···················· | ···················· | ···················· | |
| D | 91 hCoV-19/Belgium/UGent-85/2020\|EPI_ISL_468733\|2020- | ···················· | ···················· | ···················· | ···················· | ···················· | ···················· | |
| D | 92 hCoV-19/Belgium/UGent-86/2020\|EPI_ISL_468734\|2020- | ···················· | ···················· | ···················· | ···················· | ···················· | ···················· | |
| D | 93 hCoV-19/Belgium/UGent-87/2020\|EPI_ISL_468735\|2020- | ···················· | ···················· | ···················· | ···················· | ···················· | ···················· | |
| D | 94 hCoV-19/Belgium/UGent-88/2020\|EPI_ISL_468736\|2020- | ···················· | ···················· | ···················· | ···················· | ···················· | ···················· | |
| D | 95 hCoV-19/Belgium/UGent-89/2020\|EPI_ISL_468737\|2020- | ···················· | ···················· | ···················· | ···················· | ···················· | ···················· | |
| D | 96 hCoV-19/Belgium/UGent-90/2020\|EPI_ISL_468738\|2020- | ···················· | ···················· | ···················· | ···················· | ···················· | ···················· | |
| D | 97 hCoV-19/Belgium/UGent-91/2020\|EPI_ISL_468739\|2020- | ···················· | ···················· | ···················· | ···················· | ···················· | ···················· | |
| D | 98 hCoV-19/Belgium/UGent-92/2020\|EPI_ISL_468740\|2020- | ···················· | ···················· | ···················· | ···················· | ···················· | ···················· | |
| D | 99 hCoV-19/Belgium/UGent-93/2020\|EPI_ISL_468741\|2020- | ···················· | ···················· | ···················· | ···················· | ···················· | ···················· | |
| D | 100 hCoV-19/Belgium/UGent-95/2020\|EPI_ISL_468743\|2020- | ···················· | ···················· | ···················· | ···················· | ···················· | ···················· | |
| D | 101 hCoV-19/Belgium/UGent-96/2020\|EPI_ISL_468744\|2020- | ···················· | ···················· | ···················· | ···················· | ···················· | ···················· | |
| D | 102 hCoV-19/Belgium/UGent-97/2020\|EPI_ISL_468745\|2020- | ···················· | ···················· | ···················· | ···················· | ···················· | ···················· | |
| D | 103 hCoV-19/Belgium/UGent-98/2020\|EPI_ISL_468746\|2020- | ···················· | ···················· | ···················· | ···················· | ···················· | ···················· | |
| D | 104 hCoV-19/Chile/RM-CMM-05230/2020\|EPI_ISL_468747\|202 | ···················· | ···················· | ···················· | ···················· | ···················· | ···················· | |
| D | 105 hCoV-19/Chile/RM-CMM-05207/2020\|EPI_ISL_468748\|202 | ···················· | ···················· | ···················· | ···················· | ···················· | ···················· | |
| D | 106 hCoV-19/Chile/RM-CMM-04917/2020\|EPI_ISL_468749\|202 | ···················· | ···················· | ···················· | ···················· | ···················· | ···················· | |
| D | 107 hCoV-19/Chile/RM-CMM-05426/2020\|EPI_ISL_468750\|202 | ···················· | ···················· | ···················· | ···················· | ···················· | ···················· | |
| D | 108 hCoV-19/Chile/RM-CMM-05015/2020\|EPI_ISL_468751\|202 | ···················· | ···················· | ···················· | ···················· | ···················· | ···················· | |
| D | 109 hCoV-19/Chile/RM-CMM-23741/2020\|EPI_ISL_468752\|202 | ···················· | ···················· | ···················· | ···················· | ···················· | ···················· | |
| D | 110 hCoV-19/Chile/AT-CMM-00015/2020\|EPI_ISL_468757\|202 | ···················· | ···················· | ···················· | ···················· | ···················· | ···················· | |
| D | 111 hCoV-19/Chile/AT-CMM-00063/2020\|EPI_ISL_468758\|202 | ···················· | ···················· | ···················· | ···················· | ···················· | ···················· | |
| D | 112 hCoV-19/Chile/AT-CMM-00064/2020\|EPI_ISL_468759\|202 | ···················· | ···················· | ···················· | ···················· | ···················· | ···················· | |
| D | 113 hCoV-19/Chile/BI-CMM-25338/2020\|EPI_ISL_468760\|202 | ···················· | ···················· | ···················· | ···················· | ···················· | ···················· | |
| D | 114 hCoV-19/Spain/RI-IBV-h2361/2020\|EPI_ISL_468761\|202 | ···················· | ···················· | ··A················· | ···················· | ···················· | ···················· | |
| D | 115 hCoV-19/Spain/RI-IBV-h2286/2020\|EPI_ISL_468763\|202 | ···················· | ···················· | ···················· | ···················· | ···················· | ···················· | |
| D | 116 hCoV-19/Spain/RI-IBV-h2285/2020\|EPI_ISL_468764\|202 | ···················· | ···················· | ···················· | ···················· | ···················· | ···················· | |
| D | 122 hCoV-19/Spain/AR-IBV-2487/2020\|EPI_ISL_468771\|2020 | ···················· | ···················· | ···················· | ···················· | ···················· | ···················· | |
| D | 128 hCoV-19/Wales/PHWC-162DAA/2020\|EPI_ISL_473200\|2020 | ···················· | ···················· | ···················· | ···················· | ···················· | ···················· | |
| D | 129 hCoV-19/Wales/PHWC-162DD7/2020\|EPI_ISL_473202\|2020 | ···················· | ···················· | ···················· | ···················· | ···················· | ···················· | |
| D | 130 hCoV-19/Wales/PHWC-162E10/2020\|EPI_ISL_473206\|2020 | ···················· | ···················· | ···················· | ···················· | ···················· | ···················· | |
| D | 131 hCoV-19/Wales/PHWC-162E6B/2020\|EPI_ISL_473210\|2020 | ···················· | ···················· | ···················· | ···················· | ···················· | ···················· | |
| D | 132 hCoV-19/Wales/PHWC-162E7A/2020\|EPI_ISL_473211\|2020 | ···················· | ···················· | ···················· | ···················· | ···················· | ···················· | |
| D | 133 hCoV-19/Wales/PHWC-162F59/2020\|EPI_ISL_473223\|2020 | ···················· | ···················· | ···················· | ···················· | ···················· | ···················· | |
| D | 134 hCoV-19/Wales/PHWC-162FD1/2020\|EPI_ISL_473230\|2020 | ···················· | ···················· | ···················· | ···················· | ···················· | ···················· | |
| D | 135 hCoV-19/Wales/PHWC-162FE0/2020\|EPI_ISL_473231\|2020 | ···················· | ···················· | ···················· | ···················· | ···················· | ···················· | |
| D | 136 hCoV-19/Wales/PHWC-162FFF/2020\|EPI_ISL_473232\|2020 | ···················· | ···················· | ···················· | ···················· | ···················· | ···················· | |
| D | 137 hCoV-19/Wales/PHWC-163037/2020\|EPI_ISL_473236\|2020 | ···················· | ···················· | ···················· | ···················· | ···················· | ···················· | |
| D | 138 hCoV-19/Wales/PHWC-163055/2020\|EPI_ISL_473238\|2020 | ···················· | ···················A | ···················· | ···················· | ···················· | ···················· | 1 |
| D | 139 hCoV-19/Wales/PHWC-163064/2020\|EPI_ISL_473239\|2020 | ···················· | ···················· | ···················· | ···················· | ···················· | ···················· | |
| D | 140 hCoV-19/Wales/PHWC-163082/2020\|EPI_ISL_473241\|2020 | ···················· | ···················· | ···················· | ···················· | ···················· | ···················· | |
| D | 141 hCoV-19/Wales/PHWC-1630DD/2020\|EPI_ISL_473246\|2020 | ···················· | ···················· | ···················· | ···················· | ···················· | ···················· | |
| D | 142 hCoV-19/Wales/PHWC-163116/2020\|EPI_ISL_473249\|2020 | ···················· | ···················· | ···················· | ···················· | ···················· | ···················· | |
| D | 143 hCoV-19/Wales/PHWC-163125/2020\|EPI_ISL_473250\|2020 | ···················· | ···················· | ···················· | ···················· | ···················· | ···················· | |
| D | 144 hCoV-19/Wales/PHWC-163161/2020\|EPI_ISL_473254\|2020 | ···················· | ···················· | ···················· | ···················· | ···················· | ···················· | |
| D | 145 hCoV-19/Wales/PHWC-1631BC/2020\|EPI_ISL_473259\|2020 | ···················· | ···················· | ···················· | ···················· | ···················· | ···················· | |
| D | 146 hCoV-19/Wales/PHWC-1631E9/2020\|EPI_ISL_473261\|2020 | ···················· | ···················· | ···················· | ···················· | ···················· | ···················· | |
| D | 147 hCoV-19/Wales/PHWC-163231/2020\|EPI_ISL_473266\|2020 | ···················· | ···················· | ···················· | ···················· | ···················· | ···················· | |
| D | 148 hCoV-19/Wales/PHWC-16336B/2020\|EPI_ISL_473283\|2020 | ···················· | ···················· | ···················· | ···················· | ···················· | ···················· | |
| D | 149 hCoV-19/Wales/PHWC-163389/2020\|EPI_ISL_473285\|2020 | ···················· | ···················· | ···················· | ···················· | ···················· | ···················· | |
| D | 150 hCoV-19/Wales/PHWC-163398/2020\|EPI_ISL_473286\|2020 | ···················· | ···················· | ···················· | ···················· | ···················· | ···················· | |
| D | 151 hCoV-19/Wales/PHWC-1633A7/2020\|EPI_ISL_473287\|2020 | ···················· | ···················· | ···················· | ···················· | ···················· | ···················· | |
| D | 152 hCoV-19/Wales/PHWC-1633C5/2020\|EPI_ISL_473289\|2020 | ···················· | ···················· | ···················· | ···················· | ···················· | ···················· | |
| D | 153 hCoV-19/Wales/PHWC-1633E3/2020\|EPI_ISL_473291\|2020 | ···················· | ···················· | ···················· | ···················· | ···················· | ···················· | |
| D | 154 hCoV-19/Wales/PHWC-1633F2/2020\|EPI_ISL_473292\|2020 | ···················· | ···················· | ···················· | ···················· | ···················· | ···················· | |
| D | 155 hCoV-19/Wales/PHWC-16342C/2020\|EPI_ISL_473294\|2020 | ···················· | ···················· | ···················· | ···················· | ···················· | ···················· | |
| D | 156 hCoV-19/Wales/PHWC-16344A/2020\|EPI_ISL_473296\|2020 | ···················· | ···················· | ···················· | ···················· | ···················· | ···················· | |
| D | 157 hCoV-19/Wales/PHWC-163459/2020\|EPI_ISL_473297\|2020 | ···················· | ···················· | ···················· | ···················· | ···················· | ···················· | |
| D | 158 hCoV-19/Wales/PHWC-35952/2020\|EPI_ISL_474501\|2020- | ···················· | ···················· | ···················· | ···················· | ···················· | ···················· | |
| D | 159 hCoV-19/Wales/PHWC-3598F/2020\|EPI_ISL_474504\|2020- | ···················· | ···················· | ···················· | ···················· | ···················· | ···················· | |
| D | 160 hCoV-19/Wales/PHWC-359DA/2020\|EPI_ISL_474509\|2020- | ···················· | ···················· | ···················· | ···················· | ···················· | ···················· | |
| D | 161 hCoV-19/Wales/PHWC-35A5F/2020\|EPI_ISL_474517\|2020- | ···················· | ···················· | ···················· | ···················· | ···················· | ···················· | |
| D | 162 hCoV-19/Wales/PHWC-35AAA/2020\|EPI_ISL_474522\|2020- | ···················· | ···················· | ···················· | ···················· | ···················· | ···················· | |
| D | 163 hCoV-19/Wales/PHWC-35AD7/2020\|EPI_ISL_474525\|2020- | ···················· | ···················· | ···················· | ···················· | ···················· | ···················· | |
| D | 164 hCoV-19/Wales/PHWC-35AE6/2020\|EPI_ISL_474526\|2020- | ···················· | ···················· | ···················· | ···················· | ···················· | ···················· | |
| D | 165 hCoV-19/Wales/PHWC-35BA7/2020\|EPI_ISL_474537\|2020- | ···················· | ···················· | ···················· | ···················· | ···················· | ···················· | |
| D | 166 hCoV-19/Wales/PHWC-35BD4/2020\|EPI_ISL_474540\|2020- | ···················· | ···················· | ···················· | ···················· | ···················· | ···················· | |

| | | | | | | | | | |
|---|---|---|---|---|---|---|---|---|---|
| D | 167 | hCoV-19/Wales/PHWC-35BF2/2020\|EPI_ISL_474542\|2020- | ···················· | ···················· | ···················· | ···················· | ···················· | ···················· | |
| D | 168 | hCoV-19/Wales/PHWC-35C0E/2020\|EPI_ISL_474543\|2020- | ···················· | ···················· | ···················· | ···················· | ···················· | ···················· | |
| D | 169 | hCoV-19/Wales/PHWC-35C2C/2020\|EPI_ISL_474545\|2020- | ···················· | ···················· | ···················· | ···················· | ···················· | ···················· | |
| D | 170 | hCoV-19/Wales/PHWC-35C95/2020\|EPI_ISL_474551\|2020- | ···················· | ···················· | ···················· | ···················· | ···················· | ···················· | |
| D | 171 | hCoV-19/Wales/PHWC-35CB3/2020\|EPI_ISL_474553\|2020- | ···················· | ···················· | ···················· | ···················· | ···················· | ···················· | |
| D | 172 | hCoV-19/Wales/PHWC-35CE0/2020\|EPI_ISL_474556\|2020- | ···················· | ···················· | ···················· | ···················· | ···················· | ···················· | |
| D | 173 | hCoV-19/Wales/PHWC-35CFF/2020\|EPI_ISL_474557\|2020- | ···················· | ···················· | ···················· | ···················· | ···················· | ···················· | |
| D | 174 | hCoV-19/Wales/PHWC-35D83/2020\|EPI_ISL_474566\|2020- | ···················· | ···················· | ···················· | ···················· | ···················· | ···················· | |
| D | 175 | hCoV-19/Wales/PHWC-35E26/2020\|EPI_ISL_474576\|2020- | ···················· | ···················· | ···················· | ···················· | ···················· | ···················· | |
| D | 176 | hCoV-19/Wales/PHWC-35E62/2020\|EPI_ISL_474580\|2020- | ···················· | ···················· | ···················· | ···················· | ···················· | ···················· | |
| D | 177 | hCoV-19/Wales/PHWC-35EEA/2020\|EPI_ISL_474588\|2020- | ···················· | ···················· | ···················· | ···················· | ···················· | ···················· | |
| D | 178 | hCoV-19/Wales/PHWC-35F23/2020\|EPI_ISL_474592\|2020- | ···················· | ···················· | ···················· | ···················· | ···················· | ···················· | |
| D | 179 | hCoV-19/Wales/PHWC-35F50/2020\|EPI_ISL_474595\|2020- | ···················· | ···················· | ···················· | ···················· | ···················· | ···················· | |
| D | 180 | hCoV-19/Wales/PHWC-35F7E/2020\|EPI_ISL_474597\|2020- | ···················· | ···················· | ···················· | ···················· | ···················· | ···················· | |
| D | 181 | hCoV-19/Austria/CeMM0376/2020\|EPI_ISL_475820\|2020- | ···················· | ···················· | ···················· | ···················· | ···················· | ···················· | |
| D | 182 | hCoV-19/Austria/CeMM0377/2020\|EPI_ISL_475821\|2020- | ···················· | ···················· | ···················· | ···················· | ···················· | ···················· | |
| D | 183 | hCoV-19/Austria/CeMM0379/2020\|EPI_ISL_475822\|2020- | ···················· | ···················· | ···················· | ···················· | ···················· | ···················· | |
| D | 184 | hCoV-19/Austria/CeMM0380/2020\|EPI_ISL_475823\|2020- | ···················· | ···················· | ···················· | ···················· | ···················· | ···················· | |
| D | 185 | hCoV-19/Austria/CeMM0381/2020\|EPI_ISL_475824\|2020- | ···················· | ···················· | ···················· | ···················· | ···················· | ···················· | |
| D | 186 | hCoV-19/Austria/CeMM0382/2020\|EPI_ISL_475825\|2020- | ···················· | ···················· | ···················· | ···················· | ···················· | ···················· | |
| D | 187 | hCoV-19/Austria/CeMM0384/2020\|EPI_ISL_475826\|2020- | ···················· | ···················· | ···················· | ···················· | ···················· | ···················· | |
| D | 188 | hCoV-19/Austria/CeMM0385/2020\|EPI_ISL_475827\|2020- | ···················· | ···················· | ···················· | ···················· | ···················· | ···················· | |
| D | 189 | hCoV-19/Austria/CeMM0386/2020\|EPI_ISL_475828\|2020- | ···················· | ···················· | ···················· | ···················· | ···················· | ···················· | |
| D | 190 | hCoV-19/Austria/CeMM0388/2020\|EPI_ISL_475829\|2020- | ···················· | ···················· | ···················· | ···················· | ···················· | ···················· | |
| D | 191 | hCoV-19/Austria/CeMM0390/2020\|EPI_ISL_475830\|2020- | ···················· | ···················· | ···················· | ···················· | ···················· | ···················· | |
| D | 192 | hCoV-19/Austria/CeMM0391/2020\|EPI_ISL_475831\|2020- | ···················· | ···················· | ···················· | ···················· | ···················· | ···················· | |
| D | 193 | hCoV-19/Austria/CeMM0392/2020\|EPI_ISL_475832\|2020- | ···················· | ···················· | ···················· | ···················· | ···················· | ···················· | |
| D | 194 | hCoV-19/Austria/CeMM0393/2020\|EPI_ISL_475833\|2020- | ···················· | ···················· | ···················· | ···················· | ···················· | ···················· | |
| D | 195 | hCoV-19/Austria/CeMM0397/2020\|EPI_ISL_475834\|2020- | ···················· | ···················· | ···················· | ···················· | ···················· | ···················· | |
| D | 196 | hCoV-19/Austria/CeMM0400/2020\|EPI_ISL_475835\|2020- | ···················· | ···················· | ···················· | ···················· | ···················· | ···················· | |
| D | 197 | hCoV-19/Austria/CeMM0401/2020\|EPI_ISL_475836\|2020- | ···················· | ···················· | ···················· | ···················· | ···················· | ···················· | |
| D | 198 | hCoV-19/Austria/CeMM0402/2020\|EPI_ISL_475837\|2020- | ···················· | ···················· | ···················· | ···················· | ···················· | ···················· | |
| D | 199 | hCoV-19/Austria/CeMM0403/2020\|EPI_ISL_475838\|2020- | ···················· | ···················· | ···················· | ···················· | ···················· | ···················· | |
| D | 200 | hCoV-19/Austria/CeMM0404/2020\|EPI_ISL_475839\|2020- | ···················· | ···················A | ···················· | ···················· | ···················· | ···················· | 1 |
| D | 201 | hCoV-19/Austria/CeMM0408/2020\|EPI_ISL_475840\|2020- | ···················· | ···················· | ···················· | ···················· | ···················· | ···················· | |
| D | 202 | hCoV-19/Austria/CeMM0409/2020\|EPI_ISL_475841\|2020- | ···················· | ···················· | ···················· | ···················· | ···················· | ···················· | |
| D | 203 | hCoV-19/Austria/CeMM0412/2020\|EPI_ISL_475842\|2020- | ···················· | ···················· | ···················· | ···················· | ···················· | ···················· | |
| D | 204 | hCoV-19/Austria/CeMM0413/2020\|EPI_ISL_475843\|2020- | ···················· | ···················· | ···················· | ···················· | ···················· | ···················· | |
| D | 205 | hCoV-19/Austria/CeMM0414/2020\|EPI_ISL_475844\|2020- | ···················· | ···················· | ···················· | ···················· | ···················· | ···················· | |
| D | 206 | hCoV-19/Austria/CeMM0417/2020\|EPI_ISL_475845\|2020- | ···················· | ···················· | ···················· | ···················· | ···················· | ···················· | |
| D | 207 | hCoV-19/Austria/CeMM0420/2020\|EPI_ISL_475848\|2020- | ···················· | ···················· | ···················· | ···················· | ···················· | ···················· | |
| D | 208 | hCoV-19/Austria/CeMM0421/2020\|EPI_ISL_475849\|2020- | ···················· | ···················· | ···················· | ···················· | ···················· | ···················· | |
| D | 209 | hCoV-19/Austria/CeMM0422/2020\|EPI_ISL_475850\|2020- | ···················· | ···················· | ···················· | ···················· | ···················· | ···················· | |
| D | 210 | hCoV-19/Austria/CeMM0423/2020\|EPI_ISL_475851\|2020- | ···················· | ···················· | ···················· | ···················· | ···················· | ···················· | |
| D | 211 | hCoV-19/Austria/CeMM0424/2020\|EPI_ISL_475852\|2020- | ···················· | ···················· | ···················· | ···················· | ···················· | ···················· | |
| D | 212 | hCoV-19/Austria/CeMM0425/2020\|EPI_ISL_475853\|2020- | ···················· | ···················· | ···················· | ···················· | ···················· | ···················· | |
| D | 213 | hCoV-19/Austria/CeMM0426/2020\|EPI_ISL_475854\|2020- | ···················· | ···················· | ···················· | ···················· | ···················· | ···················· | |
| D | 214 | hCoV-19/Austria/CeMM0427/2020\|EPI_ISL_475855\|2020- | ···················· | ···················· | ···················· | ···················· | ···················· | ···················· | |
| D | 215 | hCoV-19/Austria/CeMM0432/2020\|EPI_ISL_475859\|2020- | ···················· | ···················· | ···················· | ···················· | ···················· | ···················· | |
| D | 216 | hCoV-19/Austria/CeMM0433/2020\|EPI_ISL_475860\|2020- | ···················· | ···················· | ···················· | ···················· | ···················· | ···················· | |
| D | 217 | hCoV-19/Austria/CeMM0435/2020\|EPI_ISL_475861\|2020- | ···················· | ···················· | ···················· | ···················· | ···················· | ···················· | |
| D | 218 | hCoV-19/Austria/CeMM0436/2020\|EPI_ISL_475862\|2020- | ···················· | ···················· | ···················· | ···················· | ···················· | ···················· | |
| D | 219 | hCoV-19/Austria/CeMM0439/2020\|EPI_ISL_475863\|2020- | ···················· | ···················· | ···················· | ···················· | ···················· | ···················· | |
| D | 220 | hCoV-19/Austria/CeMM0441/2020\|EPI_ISL_475864\|2020- | ···················· | ···················· | ···················· | ···················· | ···················· | ···················· | |
| D | 221 | hCoV-19/Austria/CeMM0448/2020\|EPI_ISL_475865\|2020- | ···················· | ···················· | ···················· | ···················· | ···················· | ···················· | |
| D | 222 | hCoV-19/Austria/CeMM0449/2020\|EPI_ISL_475866\|2020- | ···················· | ···················· | ···················· | ···················· | ···················· | ···················· | |
| D | 223 | hCoV-19/Austria/CeMM0454/2020\|EPI_ISL_475869\|2020- | ···················· | ···················· | ···················· | ···················· | ···················· | ···················· | |
| D | 224 | hCoV-19/Austria/CeMM0458/2020\|EPI_ISL_475870\|2020- | ···················· | ···················· | ···················· | ···················· | ···················· | ···················· | |
| D | 225 | hCoV-19/Austria/CeMM0460/2020\|EPI_ISL_475871\|2020- | ···················· | ···················· | ···················· | ···················· | ···················· | ···················· | |
| D | 226 | hCoV-19/Austria/CeMM0461/2020\|EPI_ISL_475872\|2020- | ···················· | ···················· | ···················· | ···················· | ···················· | ···················· | |
| D | 227 | hCoV-19/Austria/CeMM0462/2020\|EPI_ISL_475873\|2020- | ···················· | ···················· | ···················· | ···················· | ···················· | ···················· | |
| D | 228 | hCoV-19/Austria/CeMM0463/2020\|EPI_ISL_475874\|2020- | ···················· | ···················· | ···················· | ···················· | ···················· | ···················· | |
| D | 229 | hCoV-19/Austria/CeMM0466/2020\|EPI_ISL_475875\|2020- | ···················· | ···················· | ···················· | ···················· | ···················· | ···················· | |
| D | 230 | hCoV-19/Austria/CeMM0468/2020\|EPI_ISL_475876\|2020- | ···················· | ···················· | ···················· | ···················· | ···················· | ···················· | |
| D | 231 | hCoV-19/Austria/CeMM0473/2020\|EPI_ISL_475878\|2020- | ···················· | ···················· | ···················· | ···················· | ···················· | ···················· | |
| D | 232 | hCoV-19/Austria/CeMM0474/2020\|EPI_ISL_475879\|2020- | ···················· | ···················· | ···················· | ···················· | ···················· | ···················· | |
| D | 233 | hCoV-19/Austria/CeMM0475/2020\|EPI_ISL_475880\|2020- | ···················· | ···················· | ···················· | ···················· | ···················· | ···················· | |
| D | 234 | hCoV-19/Austria/CeMM0479/2020\|EPI_ISL_475882\|2020- | ···················· | ···················· | ···················· | ···················· | ···················· | ···················· | |
| D | 235 | hCoV-19/Austria/CeMM0480/2020\|EPI_ISL_475883\|2020- | ········Y··········· | ···················· | ···················· | ···················· | ···················· | ···················· | |
| D | 236 | hCoV-19/Austria/CeMM0481/2020\|EPI_ISL_475884\|2020- | ···················· | ···················· | ···················· | ···················· | ···················· | ···················· | |
| D | 237 | hCoV-19/Austria/CeMM0483/2020\|EPI_ISL_475885\|2020- | ···················· | ···················· | ···················· | ···················· | ···················· | ···················· | |
| D | 238 | hCoV-19/Austria/CeMM0484/2020\|EPI_ISL_475886\|2020- | ···················· | ···················· | ···················· | ···················· | ···················· | ···················· | |
| D | 239 | hCoV-19/Austria/CeMM0516/2020\|EPI_ISL_475889\|2020- | ···················· | ···················· | ···················· | ···················· | ···················· | ···················· | |
| D | 240 | hCoV-19/Austria/CeMM0517/2020\|EPI_ISL_475890\|2020- | ···················· | ···················· | ···················· | ···················· | ···················· | ···················· | |
| D | 241 | hCoV-19/Austria/CeMM0519/2020\|EPI_ISL_475891\|2020- | ···················· | ···················· | ···················· | ···················· | ···················· | ···················· | |
| D | 242 | hCoV-19/Austria/CeMM0521/2020\|EPI_ISL_475893\|2020- | ···················· | ···················· | ···················· | ···················· | ···················· | ···················· | |
| D | 243 | hCoV-19/Austria/CeMM0527/2020\|EPI_ISL_475894\|2020- | ···················· | ···················· | ···················· | ···················· | ···················· | ···················· | |
| D | 244 | hCoV-19/Austria/CeMM0528/2020\|EPI_ISL_475895\|2020- | ···················· | ···················· | ···················· | ···················· | ···················· | ···················· | |

```
D    245 hCoV-19/Austria/CeMM0530/2020|EPI_ISL_475896|2020-  ....................  ....................  ....................  ....................  ....................  ....................
D    246 hCoV-19/Austria/CeMM0532/2020|EPI_ISL_475897|2020-  ....................  ....................  ....................  ....................  ....................  ....................
D    247 hCoV-19/Austria/CeMM0536/2020|EPI_ISL_475899|2020-  ....................  ....................  ....................  ....................  ....................  ....................
D    248 hCoV-19/Scotland/CVR3450/2020|EPI_ISL_478060|2020-  ....................  ....................  ....................  ....................  ....................  ....................
D    249 hCoV-19/Scotland/CVR3454/2020|EPI_ISL_478062|2020-  ....................  ....................  ....................  ....................  ....................  ....................
D    251 hCoV-19/Scotland/CVR3464/2020|EPI_ISL_478068|2020-  ....................  ...................A  ....................  ....................  ....................  ....................  1
D    252 hCoV-19/Wales/PHWC-164F0C/2020|EPI_ISL_479397|2020  ....................  ....................  ....................  ....................  ....................  ....................
D    253 hCoV-19/Wales/PHWC-164F75/2020|EPI_ISL_479403|2020  ....................  ....................  ....................  ....................  ....................  ....................
D    254 hCoV-19/Wales/PHWC-164FEE/2020|EPI_ISL_479410|2020  ....................  ....................  ....................  ....................  ....................  ....................
D    255 hCoV-19/Wales/PHWC-165026/2020|EPI_ISL_479412|2020  ....................  ....................  ....................  ....................  ....................  ....................
D    256 hCoV-19/Wales/PHWC-1650CC/2020|EPI_ISL_479420|2020  ....................  ....................  ....................  ....................  ....................  ....................
D    257 hCoV-19/England/20144037904/2020|EPI_ISL_466000|20  ....................  ....................  ....................  ....................  ....................  ....................
D    258 hCoV-19/England/20144050404/2020|EPI_ISL_466001|20  ....................  ....................  ....................  ....................  ....................  ....................
D    259 hCoV-19/England/20144052804/2020|EPI_ISL_466002|20  ....................  ....................  ....................  ....................  ....................  ....................
D    260 hCoV-19/England/20144057404/2020|EPI_ISL_466003|20  ....................  ....................  ....................  ....................  ....................  ....................
D    263 hCoV-19/England/20144067704/2020|EPI_ISL_466006|20  ....................  ....................  ....................  ....................  ....................  ....................
D    265 hCoV-19/England/20146003804/2020|EPI_ISL_466014|20  ....................  ....................  ....................  ....................  ....................  ....................
D    266 hCoV-19/England/20146003904/2020|EPI_ISL_466015|20  ....................  ....................  ....................  ....................  ....................  ....................
D    267 hCoV-19/England/20146005604/2020|EPI_ISL_466016|20  ....................  ....................  ....................  ....................  ....................  ....................
D    268 hCoV-19/England/20146005904/2020|EPI_ISL_466017|20  ....................  ....................  ....................  ....................  ....................  ....................
D    269 hCoV-19/England/20146006804/2020|EPI_ISL_466018|20  ....................  ....................  ....................  ....................  ....................  ....................
D    270 hCoV-19/England/20146007804/2020|EPI_ISL_466023|20  ....................  ....................  ....................  ....................  ....................  ....................
D    271 hCoV-19/England/20146008604/2020|EPI_ISL_466025|20  ....................  ....................  ....................  ....................  ....................  ....................
D    272 hCoV-19/England/20146010404/2020|EPI_ISL_466026|20  ....................  ....................  ....................  ....................  ....................  ....................
D    273 hCoV-19/England/20146011404/2020|EPI_ISL_466028|20  ....................  ....................  ....................  ....................  ....................  ....................
D    275 hCoV-19/England/20146013904/2020|EPI_ISL_466030|20  ....................  ....................  ....................  ....................  ....................  ....................
D    276 hCoV-19/England/20146015604/2020|EPI_ISL_466032|20  ....................  ....................  ....................  ....................  ....................  ....................
D    277 hCoV-19/England/20146015904/2020|EPI_ISL_466033|20  ....................  ....................  ....................  ....................  ....................  ....................
D    278 hCoV-19/England/20146016004/2020|EPI_ISL_466034|20  ....................  ....................  ....................  ....................  ....................  ....................
D    279 hCoV-19/England/20146018204/2020|EPI_ISL_466036|20  ....................  ....................  ....................  ....................  ....................  ....................
D    280 hCoV-19/England/20146018302/2020|EPI_ISL_466037|20  ....................  ....................  ....................  ....................  ....................  ....................
D    281 hCoV-19/England/20146019504/2020|EPI_ISL_466038|20  ....................  ....................  ....................  ....................  ....................  ....................
D    282 hCoV-19/England/20146019804/2020|EPI_ISL_466039|20  ....................  ....................  ....................  ....................  ....................  ....................
D    283 hCoV-19/England/20146021204/2020|EPI_ISL_466040|20  ....................  ....................  ....................  ....................  ....................  ....................
D    284 hCoV-19/England/20146021304/2020|EPI_ISL_466041|20  ....................  ....................  ....................  ....................  ....................  ....................
D    285 hCoV-19/England/20146057204/2020|EPI_ISL_466043|20  ....................  ....................  ....................  ....................  ....................  ....................
D    286 hCoV-19/England/20146057704/2020|EPI_ISL_466044|20  ....................  ....................  ....................  ....................  ....................  ....................
D    288 hCoV-19/England/20146064004/2020|EPI_ISL_466046|20  ....................  ....................  ....................  ....................  ....................  ....................
D    289 hCoV-19/England/20146064204/2020|EPI_ISL_466047|20  ....................  ....................  ....................  ....................  ....................  ....................
D    290 hCoV-19/England/20148001404/2020|EPI_ISL_466048|20  ....................  ....................  ....................  ....................  ....................  ....................
D    293 hCoV-19/England/20148006204/2020|EPI_ISL_466052|20  ....................  ....................  ....................  ....................  ....................  ....................
D    294 hCoV-19/England/20148032904/2020|EPI_ISL_466054|20  ....................  ....................  ....................  ....................  ....................  ....................
D    297 hCoV-19/England/20148035204/2020|EPI_ISL_466061|20  ....................  ....................  ....................  ....................  ....................  ....................
D    298 hCoV-19/England/20148035404/2020|EPI_ISL_466062|20  ....................  ....................  ....................  ....................  ....................  ....................
D    300 hCoV-19/England/20148036404/2020|EPI_ISL_466065|20  ....................  ....................  ....................  ....................  ....................  ....................
D    302 hCoV-19/England/20148041804/2020|EPI_ISL_466068|20  ....................  ....................  ....................  ....................  ....................  ....................
D    303 hCoV-19/England/20148045404/2020|EPI_ISL_466069|20  ....................  ....................  ....................  ....................  ....................  ....................
D    304 hCoV-19/England/20148049704/2020|EPI_ISL_466071|20  ....................  ....................  ....................  ....................  ....................  ....................
D    305 hCoV-19/England/20148066804/2020|EPI_ISL_466074|20  ....................  ....................  ....................  ....................  ....................  ....................
D    307 hCoV-19/England/20148067204/2020|EPI_ISL_466076|20  ....................  ....................  ....................  ....................  ....................  ....................
D    308 hCoV-19/England/20148067304/2020|EPI_ISL_466077|20  ....................  ....................  ....................  ....................  ....................  ....................
D    309 hCoV-19/Canada/BC_11656063/2020|EPI_ISL_467327|202  ....................  .A..................  ....................  ....................  ....................  ....................
D    310 hCoV-19/Canada/BC_11726488/2020|EPI_ISL_467329|202  ....................  ....................  ....................  ....................  ....................  ....................
D    311 hCoV-19/Canada/BC_12186719/2020|EPI_ISL_467330|202  ....................  ....................  ....................  ....................  ....................  ....................
D    312 hCoV-19/Canada/BC_12199086/2020|EPI_ISL_467331|202  ....................  ....................  ....................  ....................  ....................  ....................
D    313 hCoV-19/Canada/BC_12210980/2020|EPI_ISL_467332|202  ....................  ....................  ....................  ....................  ....................  ....................
D    314 hCoV-19/Canada/BC_12246693/2020|EPI_ISL_467333|202  ....................  ....................  ....................  ....................  ....................  ....................
D    315 hCoV-19/Canada/BC_12251717/2020|EPI_ISL_467334|202  ....................  ....................  ....................  ....................  ....................  ....................
D    316 hCoV-19/Canada/BC_12477112/2020|EPI_ISL_467335|202  ....................  ....................  ....................  ....................  ....................  ....................
D    317 hCoV-19/Canada/BC_12661418/2020|EPI_ISL_467336|202  ....................  ....................  ....................  ....................  ....................  ....................
D    318 hCoV-19/Canada/BC_12775269/2020|EPI_ISL_467337|202  ....................  ....................  ....................  ....................  ....................  ....................
D    319 hCoV-19/Canada/BC_12793935/2020|EPI_ISL_467338|202  ....................  ....................  ....................  ....................  ....................  ....................
D    320 hCoV-19/Canada/BC_13086747/2020|EPI_ISL_467339|202  ....................  ....................  ....................  ....................  ....................  ....................
D    321 hCoV-19/Canada/BC_13088259/2020|EPI_ISL_467340|202  ....................  ....................  ....................  ....................  ....................  ....................
D    322 hCoV-19/Canada/BC_13610520/2020|EPI_ISL_467341|202  ....................  ....................  ....................  ....................  ....................  ....................
D    323 hCoV-19/Canada/BC_13662361/2020|EPI_ISL_467342|202  ....................  ....................  ....................  ....................  ....................  ....................
D    324 hCoV-19/Canada/BC_13675191/2020|EPI_ISL_467343|202  ....................  ....................  ....................  ....................  ....................  ....................
D    325 hCoV-19/Brazil/RJ-1555/2020|EPI_ISL_467344|2020-04  ....................  ....................  ....................  ....................  ....................  ....................
D    326 hCoV-19/Brazil/RJ-1574/2020|EPI_ISL_467345|2020-04  ....................  ....................  ....................  ....................  ....................  ....................
D    327 hCoV-19/Brazil/RJ-1595/2020|EPI_ISL_467346|2020-04  ....................  ....................  ....................  ....................  ....................  ....................
D    328 hCoV-19/Brazil/RJ-2091/2020|EPI_ISL_467347|2020-04  ....................  ....................  ....................  ....................  ....................  ....................
D    329 hCoV-19/Brazil/RJ-2195/2020|EPI_ISL_467348|2020-04  ....................  ....................  ....................  ....................  ....................  ....................
D    330 hCoV-19/Brazil/RJ-2197/2020|EPI_ISL_467349|2020-04  ....................  ....................  ....................  ....................  ....................  ....................
D    331 hCoV-19/Brazil/RJ-2208/2020|EPI_ISL_467350|2020-04  ....................  ....................  ....................  ....................  ....................  ....................
D    332 hCoV-19/Brazil/RJ-2233/2020|EPI_ISL_467351|2020-04  ....................  ....................  ....................  ....................  ....................  ....................
D    333 hCoV-19/Brazil/RJ-2422/2020|EPI_ISL_467352|2020-04  ....................  ....................  ....................  ....................  ....................  ....................
D    334 hCoV-19/Brazil/RJ-2669/2020|EPI_ISL_467353|2020-04  ....................  ....................  ....................  ....................  ....................  ....................
D    335 hCoV-19/Brazil/RJ-2676/2020|EPI_ISL_467354|2020-04  ....................  ....................  ....................  ....................  ....................  ....................
```

| | | | | | | | |
|---|---|---|---|---|---|---|---|
| D | 336 hCoV-19/Brazil/RJ-2678/2020\|EPI_ISL_467355\|2020-04 | ···················· | ···················· | ···················· | ···················· | ···················· | ···················· |
| D | 337 hCoV-19/Brazil/RJ-2682/2020\|EPI_ISL_467356\|2020-04 | ···················· | ···················· | ···················· | ···················· | ···················· | ···················· |
| D | 338 hCoV-19/Brazil/RJ-2683/2020\|EPI_ISL_467357\|2020-04 | ···················· | ···················· | ···················· | ···················· | ···················· | ···················· |
| D | 339 hCoV-19/Brazil/RJ-2696/2020\|EPI_ISL_467358\|2020-04 | ···················· | ···················· | ···················· | ···················· | ···················· | ···················· |
| D | 340 hCoV-19/Brazil/RJ-2717/2020\|EPI_ISL_467359\|2020-04 | ···················· | ···················· | ···················· | ···················· | ···················· | ···················· |
| D | 341 hCoV-19/Brazil/RJ-2733/2020\|EPI_ISL_467360\|2020-04 | ···················· | ···················· | ···················· | ···················· | ···················· | ···················· |
| D | 342 hCoV-19/Brazil/RJ-2769/2020\|EPI_ISL_467361\|2020-04 | ···················· | ···················· | ···················· | ···················· | ···················· | ···················· |
| D | 343 hCoV-19/Brazil/RJ-2770/2020\|EPI_ISL_467362\|2020-04 | ···················· | ···················· | ···················· | ···················· | ···················· | ···················· |
| D | 344 hCoV-19/Brazil/RJ-2776/2020\|EPI_ISL_467363\|2020-04 | ···················· | ···················· | ···················· | ···················· | ···················· | ···················· |
| D | 345 hCoV-19/Brazil/RJ-2777/2020\|EPI_ISL_467364\|2020-04 | ···················· | ···················· | ···················· | ···················· | ···················· | ···················· |
| D | 346 hCoV-19/Brazil/RJ-2811/2020\|EPI_ISL_467365\|2020-04 | ···················· | ···················· | ···················· | ···················· | ···················· | ···················· |
| D | 347 hCoV-19/Brazil/RJ-2812/2020\|EPI_ISL_467366\|2020-04 | ···················· | ···················· | ···················· | ···················· | ···················· | ···················· |
| D | 348 hCoV-19/Brazil/RJ-2822/2020\|EPI_ISL_467367\|2020-04 | ···················· | ···················· | ···················· | ···················· | ···················· | ···················· |
| D | 349 hCoV-19/Brazil/RJ-2840/2020\|EPI_ISL_467368\|2020-04 | ···················· | ···················· | ···················· | ···················· | ···················· | ···················· |
| D | 350 hCoV-19/Brazil/RJ-2844/2020\|EPI_ISL_467369\|2020-04 | ···················· | ···················· | ···················· | ···················· | ···················· | ···················· |
| D | 351 hCoV-19/Brazil/RJ-2847/2020\|EPI_ISL_467370\|2020-04 | ···················· | ···················· | ···················· | ···················· | ···················· | ···················· |
| D | 352 hCoV-19/Brazil/RJ-2868/2020\|EPI_ISL_467371\|2020-04 | ···················· | ···················· | ···················· | ···················· | ···················· | ···················· |
| D | 353 hCoV-19/USA/AZ-ASU2943/2020\|EPI_ISL_467372\|2020-03 | ···················· | ···················· | ···················· | ···················· | ···················· | ···················· |
| D | 354 hCoV-19/USA/AZ-ASU2949/2020\|EPI_ISL_467373\|2020-03 | ···················· | ···················· | ···················· | ···················· | ···················· | ···················· |
| D | 355 hCoV-19/Indonesia/KI-EIJK-05/2020\|EPI_ISL_467374\|2 | ···················· | ···················· | ···················· | ···················· | ···················· | ···················· |
| D | 356 hCoV-19/Indonesia/SA-EIJK-06/2020\|EPI_ISL_467375\|2 | ···················· | ···················· | ···················· | ···················· | ···················· | ···················· |
| D | 357 hCoV-19/Indonesia/JK-EIJK-07/2020\|EPI_ISL_467376\|2 | ···················· | ···················· | ···················· | ···················· | ···················· | ···················· |
| D | 358 hCoV-19/USA/NY-NYUMC884/2020\|EPI_ISL_467377\|2020-0 | ···················· | ···················· | ···················· | ···················· | ···················· | ···················· |
| D | 359 hCoV-19/USA/NY-NYUMC885/2020\|EPI_ISL_467378\|2020-0 | ···················· | ···················· | ···················· | ···················· | ···················· | ···················· |
| D | 360 hCoV-19/USA/NY-NYUMC886/2020\|EPI_ISL_467379\|2020-0 | ···················· | ···················· | ···················· | ···················· | ···················· | ···················· |
| D | 361 hCoV-19/USA/NY-NYUMC887/2020\|EPI_ISL_467380\|2020-0 | ···················· | ···················· | ···················· | ···················· | ···················· | ···················· |
| D | 362 hCoV-19/USA/NY-NYUMC888/2020\|EPI_ISL_467381\|2020-0 | ···················· | ···················· | ···················· | ···················· | ···················· | ···················· |
| D | 363 hCoV-19/USA/NY-NYUMC891/2020\|EPI_ISL_467384\|2020-0 | ···················· | ···················· | ···················· | ···················· | ···················· | ···················· |
| D | 364 hCoV-19/USA/NY-NYUMC892/2020\|EPI_ISL_467385\|2020-0 | ···················· | ···················· | ···················· | ···················· | ···················· | ···················· |
| D | 365 hCoV-19/USA/NY-NYUMC894/2020\|EPI_ISL_467387\|2020-0 | ···················· | ···················· | ···················· | ···················· | ···················· | ···················· |
| D | 366 hCoV-19/USA/NY-NYUMC898/2020\|EPI_ISL_467391\|2020-0 | ···················· | ···················· | ···················· | ···················· | ···················· | ···················· |
| D | 367 hCoV-19/USA/NY-NYUMC901/2020\|EPI_ISL_467394\|2020-0 | ···················· | ···················· | ···················· | ···················· | ···················· | ···················· |
| D | 368 hCoV-19/USA/NY-NYUMC902/2020\|EPI_ISL_467395\|2020-0 | ···················· | ···················· | ···················· | ···················· | ···················· | ···················· |
| D | 369 hCoV-19/USA/NY-NYUMC903/2020\|EPI_ISL_467396\|2020-0 | ···················· | ···················· | ···················· | ···················· | ···················· | ···················· |
| D | 370 hCoV-19/USA/NY-NYUMC905/2020\|EPI_ISL_467398\|2020-0 | ···················· | ···················· | ···················· | ···················· | ···················· | ···················· |
| D | 371 hCoV-19/USA/NY-NYUMC906/2020\|EPI_ISL_467399\|2020-0 | ···················· | ···················· | ···················· | ···················· | ···················· | ···················· |
| D | 373 hCoV-19/South Africa/KRISP-0171/2020\|EPI_ISL_46752 | ···················· | ···················· | ···················· | ···················· | ···················· | ···················· |
| D | 374 hCoV-19/South Africa/KRISP-0172/2020\|EPI_ISL_46752 | ···················· | ···················· | ···················· | ···················· | ···················· | ···················· |
| D | 375 hCoV-19/South Africa/KRISP-0173/2020\|EPI_ISL_46752 | ···················· | ···················· | ···················· | ···················· | ···················· | ···················· |
| D | 376 hCoV-19/Canada/BC_14667611/2020\|EPI_ISL_468658\|202 | ···················· | ···················· | ···················· | ···················· | ···················· | ···················· |
| D | 377 hCoV-19/Canada/BC_14727596/2020\|EPI_ISL_468659\|202 | ···················· | ···················· | ···················· | ···················· | ···················· | ···················· |
| D | 378 hCoV-19/Canada/BC_14739124/2020\|EPI_ISL_468660\|202 | ···················· | ···················· | ···················· | ···················· | ···················· | ···················· |
| D | 379 hCoV-19/Canada/BC_14763711/2020\|EPI_ISL_468661\|202 | ···················· | ···················· | ···················· | ···················· | ···················· | ···················· |
| D | 380 hCoV-19/Canada/BC_15188754/2020\|EPI_ISL_468662\|202 | ···················· | ···················· | ···················· | ···················· | ···················· | ···················· |
| D | 381 hCoV-19/Canada/BC_15222281/2020\|EPI_ISL_468664\|202 | ···················· | ···················· | ···················· | ···················· | ···················· | ···················· |
| D | 382 hCoV-19/Canada/BC_15222496/2020\|EPI_ISL_468665\|202 | ···················· | ···················· | ···················· | ···················· | ···················· | ···················· |
| D | 383 hCoV-19/Canada/BC_15443411/2020\|EPI_ISL_468666\|202 | ···················· | ···················· | ···················· | ···················· | ···················· | ···················· |
| D | 384 hCoV-19/Canada/BC_15467213/2020\|EPI_ISL_468668\|202 | ···················· | ···················· | ···················· | ···················· | ···················· | ···················· |
| D | 385 hCoV-19/Canada/BC_15474127/2020\|EPI_ISL_468669\|202 | ···················· | ···················· | ···················· | ···················· | ···················· | ···················· |
| D | 386 hCoV-19/Canada/BC_15478567/2020\|EPI_ISL_468670\|202 | ···················· | ···················· | ···················· | ···················· | ···················· | ···················· |
| D | 387 hCoV-19/Canada/BC_15550440/2020\|EPI_ISL_468671\|202 | ···················· | ···················· | ···················· | ···················· | ···················· | ···················· |
| D | 388 hCoV-19/Canada/BC_15678040/2020\|EPI_ISL_468672\|202 | ···················· | ···················· | ···················· | ···················· | ···················· | ···················· |
| D | 389 hCoV-19/Canada/BC_15682745/2020\|EPI_ISL_468673\|202 | ···················· | ···················· | ···················· | ···················· | ···················· | ···················· |
| D | 390 hCoV-19/Canada/BC_15708725/2020\|EPI_ISL_468674\|202 | ···················· | ···················· | ···················· | ···················· | ···················· | ···················· |
| D | 391 hCoV-19/Canada/BC_15710616/2020\|EPI_ISL_468675\|202 | ···················· | ···················· | ···················· | ···················· | ···················· | ···················· |
| D | 392 hCoV-19/Canada/BC_15804748/2020\|EPI_ISL_468676\|202 | ···················· | ···················· | ···················· | ···················· | ···················· | ···················· |
| D | 393 hCoV-19/Canada/BC_15820631/2020\|EPI_ISL_468677\|202 | ···················· | ···················· | ···················· | ···················· | ···················· | ···················· |
| D | 394 hCoV-19/Canada/BC_15918444/2020\|EPI_ISL_468678\|202 | ···················· | ···················· | ···················· | ···················· | ···················· | ···················· |
| D | 395 hCoV-19/Canada/BC_16223737/2020\|EPI_ISL_468679\|202 | ···················· | ···················· | ···················· | ···················· | ···················· | ···················· |
| D | 396 hCoV-19/Canada/BC_16537755/2020\|EPI_ISL_468680\|202 | ···················· | ···················· | ···················· | ···················· | ···················· | ···················· |
| D | 397 hCoV-19/Canada/BC_16588829/2020\|EPI_ISL_468683\|202 | ···················· | ···················· | ···················· | ···················· | ···················· | ···················· |
| D | 398 hCoV-19/Canada/BC_16839757/2020\|EPI_ISL_468684\|202 | ···················· | ···················· | ···················· | ···················· | ···················· | ···················· |
| D | 399 hCoV-19/Canada/BC_16917931/2020\|EPI_ISL_468685\|202 | ···················· | ···················· | ···················· | ···················· | ···················· | ···················· |
| D | 400 hCoV-19/Canada/BC_16943416/2020\|EPI_ISL_468686\|202 | ···················· | ···················· | ···················· | ···················· | ···················· | ···················· |
| D | 401 hCoV-19/Canada/BC_16976214/2020\|EPI_ISL_468687\|202 | ···················· | ···················· | ···················· | ···················· | ···················· | ···················· |
| D | 402 hCoV-19/Canada/BC_17080413/2020\|EPI_ISL_468688\|202 | ···················· | ···················· | ···················· | ···················· | ···················· | ···················· |
| D | 403 hCoV-19/Canada/BC_17607523/2020\|EPI_ISL_468689\|202 | ···················· | ···················· | ···················· | ···················· | ···················· | ···················· |
| D | 404 hCoV-19/Canada/BC_17754491/2020\|EPI_ISL_468691\|202 | ···················· | ···················· | ···················· | ···················· | ···················· | ···················· |
| D | 405 hCoV-19/Canada/BC_17906513/2020\|EPI_ISL_468693\|202 | ···················· | ···················· | ···················· | ···················· | ···················· | ···················· |
| D | 406 hCoV-19/Canada/BC_17943129/2020\|EPI_ISL_468694\|202 | ···················· | ···················· | ···················· | ···················· | ···················· | ···················· |
| D | 407 hCoV-19/Canada/BC_18101737/2020\|EPI_ISL_468695\|202 | ···················· | ···················· | ···················· | ···················· | ···················· | ···················· |
| D | 408 hCoV-19/Canada/BC_18115071/2020\|EPI_ISL_468696\|202 | ···················· | ···················· | ···················· | ···················· | ···················· | ···················· |
| D | 409 hCoV-19/Canada/BC_18116391/2020\|EPI_ISL_468698\|202 | ···················· | ···················· | ···················· | ···················· | ···················· | ···················· |
| D | 410 hCoV-19/Canada/BC_18141519/2020\|EPI_ISL_468699\|202 | ···················· | ···················· | ···················· | ···················· | ···················· | ···················· |
| D | 416 hCoV-19/Scotland/GCVR-1721BA/2020\|EPI_ISL_469988\|2 | ···················· | ···················· | ···················· | ···················· | ···················· | ···················· |
| D | 417 hCoV-19/Scotland/GCVR-1722A8/2020\|EPI_ISL_469989\|2 | ···················· | ···················· | ···················· | ···················· | ···················· | ···················· |
| D | 418 hCoV-19/Scotland/GCVR-172026/2020\|EPI_ISL_469990\|2 | ···················· | ···················· | ···················· | ···················· | ···················· | ···················· |
| D | 419 hCoV-19/Scotland/GCVR-17217E/2020\|EPI_ISL_469991\|2 | ···················· | ·A·················· | ···················· | ···················· | ···················· | ···················· |

| | | | | | | | | |
|---|---|---|---|---|---|---|---|---|
| D | 420 hCoV-19/Scotland/GCVR-17228A/2020\|EPI_ISL_469993\|2 | .................... | .A.................. | .................... | .................... | .................... | .................... | |
| D | 421 hCoV-19/Scotland/GCVR-1721D8/2020\|EPI_ISL_469994\|2 | .................... | .................... | .................... | .................... | .................... | .................... | |
| D | 422 hCoV-19/Scotland/GCVR-172105/2020\|EPI_ISL_469997\|2 | .................... | .................... | .................... | .................... | .................... | .................... | |
| D | 423 hCoV-19/Scotland/GCVR-172017/2020\|EPI_ISL_469999\|2 | .................... | .................... | .................... | .................... | .................... | .................... | |
| D | 424 hCoV-19/Austria/CeMM0337/2020\|EPI_ISL_475800\|2020- | .................... | .................... | .................... | .................... | .................... | .................... | |
| D | 425 hCoV-19/Austria/CeMM0340/2020\|EPI_ISL_475801\|2020- | .................... | .................... | .................... | .................... | .................... | .................... | |
| D | 426 hCoV-19/Austria/CeMM0341/2020\|EPI_ISL_475802\|2020- | .................... | .................... | .................... | .................... | .................... | .................... | |
| D | 427 hCoV-19/Austria/CeMM0342/2020\|EPI_ISL_475803\|2020- | .................... | .................... | .................... | .................... | .................... | .................... | |
| D | 428 hCoV-19/Austria/CeMM0343/2020\|EPI_ISL_475804\|2020- | .................... | .................... | .................... | .................... | .................... | .................... | |
| D | 429 hCoV-19/Austria/CeMM0344/2020\|EPI_ISL_475805\|2020- | .................... | .................... | .................... | .................... | .................... | .................... | |
| D | 430 hCoV-19/Austria/CeMM0346/2020\|EPI_ISL_475806\|2020- | .................... | .................... | .................... | .................... | .................... | .................... | |
| D | 431 hCoV-19/Austria/CeMM0349/2020\|EPI_ISL_475808\|2020- | .................... | .................... | .................... | .................... | .................... | .................... | |
| D | 432 hCoV-19/Austria/CeMM0350/2020\|EPI_ISL_475809\|2020- | .................... | .................... | .................... | .................... | .................... | .................... | |
| D | 433 hCoV-19/Austria/CeMM0351/2020\|EPI_ISL_475810\|2020- | .................... | .................... | .................... | .................... | .................... | .................... | |
| D | 434 hCoV-19/Austria/CeMM0352/2020\|EPI_ISL_475811\|2020- | .................... | .................... | .................... | .................... | .................... | .................... | |
| D | 435 hCoV-19/Austria/CeMM0355/2020\|EPI_ISL_475812\|2020- | .................... | .................... | .................... | .................... | .................... | .................... | |
| D | 436 hCoV-19/Austria/CeMM0366/2020\|EPI_ISL_475813\|2020- | .................... | .................... | .................... | .................... | .................... | .................... | |
| D | 437 hCoV-19/Austria/CeMM0367/2020\|EPI_ISL_475814\|2020- | .................... | .................... | .................... | .................... | .................... | .................... | |
| D | 438 hCoV-19/Austria/CeMM0368/2020\|EPI_ISL_475815\|2020- | .................... | .................... | .................... | .................... | .................... | .................... | |
| D | 439 hCoV-19/Austria/CeMM0371/2020\|EPI_ISL_475816\|2020- | .................... | .................... | .................... | .................... | .................... | .................... | |
| D | 440 hCoV-19/Austria/CeMM0372/2020\|EPI_ISL_475817\|2020- | .................... | .................... | .................... | .................... | .................... | .................... | |
| D | 441 hCoV-19/Austria/CeMM0373/2020\|EPI_ISL_475818\|2020- | .................... | .................... | .................... | .................... | .................... | .................... | |
| D | 442 hCoV-19/Austria/CeMM0375/2020\|EPI_ISL_475819\|2020- | .................... | .................... | .................... | .................... | .................... | .................... | |
| D | 443 hCoV-19/Scotland/CVR2640/2020\|EPI_ISL_478000\|2020- | .................... | .................... | .................... | .................... | .................... | .................... | |
| D | 444 hCoV-19/Scotland/CVR2645/2020\|EPI_ISL_478001\|2020- | .................... | .................... | .................... | .................... | .................... | .................... | |
| D | 445 hCoV-19/Scotland/CVR2646/2020\|EPI_ISL_478002\|2020- | .................... | .................... | .................... | .................... | .................... | .................... | |
| D | 446 hCoV-19/Scotland/CVR2649/2020\|EPI_ISL_478003\|2020- | .................... | .................... | .................... | .................... | .................... | .................... | |
| D | 447 hCoV-19/Scotland/CVR2650/2020\|EPI_ISL_478004\|2020- | .................... | .................... | .................... | .................... | .................... | .................... | |
| D | 448 hCoV-19/Scotland/CVR2653/2020\|EPI_ISL_478005\|2020- | .................... | ...................A | .................... | .................... | .................... | .................... | 1 |
| D | 449 hCoV-19/Scotland/CVR2656/2020\|EPI_ISL_478006\|2020- | .................... | ..............C..... | .................... | .................... | .................... | .................... | |
| D | 450 hCoV-19/Scotland/CVR2658/2020\|EPI_ISL_478008\|2020- | .................... | .................... | .................... | .................... | .................... | .................... | |
| D | 451 hCoV-19/Scotland/CVR2660/2020\|EPI_ISL_478009\|2020- | .................... | .................... | .................... | .................... | .................... | .................... | |
| D | 452 hCoV-19/Scotland/CVR2661/2020\|EPI_ISL_478010\|2020- | .................... | .................... | .................... | .................... | .................... | .................... | |
| D | 453 hCoV-19/Scotland/CVR2662/2020\|EPI_ISL_478011\|2020- | .................... | .................... | .................... | .................... | .................... | .................... | |
| D | 454 hCoV-19/Scotland/CVR2664/2020\|EPI_ISL_478012\|2020- | .................... | .A.................. | .................... | .................... | .................... | .................... | |
| D | 455 hCoV-19/Scotland/CVR2668/2020\|EPI_ISL_478013\|2020- | .................... | .A.................. | .................... | .................... | .................... | .................... | |
| D | 456 hCoV-19/Scotland/CVR2669/2020\|EPI_ISL_478014\|2020- | .................... | .................... | .................... | .................... | .................... | .................... | |
| D | 457 hCoV-19/Scotland/CVR2675/2020\|EPI_ISL_478015\|2020- | .................... | .A.................. | .................... | .................... | .................... | .................... | |
| D | 458 hCoV-19/Scotland/CVR2680/2020\|EPI_ISL_478016\|2020- | .................... | .................... | .................... | .................... | .................... | .................... | |
| D | 459 hCoV-19/Scotland/CVR2681/2020\|EPI_ISL_478017\|2020- | .................... | .................... | .................... | .................... | .................... | .................... | |
| D | 460 hCoV-19/Scotland/CVR2688/2020\|EPI_ISL_478018\|2020- | .................... | ...................A | .................... | .................... | .................... | .................... | 1 |
| D | 461 hCoV-19/Scotland/CVR2692/2020\|EPI_ISL_478019\|2020- | .................... | .................... | .................... | .................... | .................... | .................... | |
| D | 462 hCoV-19/Scotland/CVR2698/2020\|EPI_ISL_478020\|2020- | .................... | .................... | .................... | .................... | .................... | .................... | |
| D | 463 hCoV-19/Scotland/CVR2700/2020\|EPI_ISL_478022\|2020- | .................... | .................... | .................... | .................... | .................... | .................... | |
| D | 464 hCoV-19/Scotland/CVR2701/2020\|EPI_ISL_478023\|2020- | .................... | .................... | .................... | .................... | .................... | .................... | |
| D | 465 hCoV-19/Scotland/CVR2704/2020\|EPI_ISL_478024\|2020- | .................... | .................... | .................... | .................... | .................... | .................... | |
| D | 466 hCoV-19/Scotland/CVR2708/2020\|EPI_ISL_478025\|2020- | .................... | ...................A | .................... | .................... | .................... | .................... | 1 |
| D | 467 hCoV-19/Scotland/CVR2728/2020\|EPI_ISL_478027\|2020- | .................... | ...................A | .................... | .................... | .................... | .................... | 1 |
| D | 468 hCoV-19/Scotland/CVR2752/2020\|EPI_ISL_478028\|2020- | .................... | ...................A | .................... | .................... | .................... | .................... | 1 |
| D | 469 hCoV-19/Scotland/CVR2756/2020\|EPI_ISL_478029\|2020- | ..................T. | ...................A | .................... | .................... | .................... | .................... | 1 |
| D | 470 hCoV-19/Scotland/CVR2758/2020\|EPI_ISL_478030\|2020- | .................... | ...................A | .................... | .................... | .................... | .................... | 1 |
| D | 471 hCoV-19/Scotland/CVR2759/2020\|EPI_ISL_478031\|2020- | .................... | .................... | .................... | .................... | .................... | .................... | |
| D | 472 hCoV-19/Scotland/CVR2763/2020\|EPI_ISL_478032\|2020- | .................... | .................... | .................... | .................... | .................... | .................... | |
| D | 473 hCoV-19/Scotland/CVR2775/2020\|EPI_ISL_478035\|2020- | .................... | .A.................. | .................... | .................... | .................... | .................... | |
| D | 474 hCoV-19/Scotland/CVR2777/2020\|EPI_ISL_478036\|2020- | .................... | .................... | .................... | .................... | .................... | .................... | |
| D | 475 hCoV-19/Scotland/CVR2829/2020\|EPI_ISL_478039\|2020- | .................... | .................... | .................... | .................... | .................... | .................... | |
| D | 476 hCoV-19/Scotland/CVR2926/2020\|EPI_ISL_478045\|2020- | .................... | ..............C..... | .................... | .................... | .................... | .................... | |
| D | 477 hCoV-19/Scotland/CVR2946/2020\|EPI_ISL_478047\|2020- | .................... | ..............C..... | .................... | .................... | .................... | .................... | |
| D | 478 hCoV-19/Scotland/CVR2949/2020\|EPI_ISL_478048\|2020- | .................... | ...................A | .................... | .................... | .................... | .................... | 1 |
| D | 479 hCoV-19/Scotland/CVR2952/2020\|EPI_ISL_478049\|2020- | .................... | .................... | .................... | .................... | .................... | .................... | |
| D | 480 hCoV-19/Scotland/CVR2955/2020\|EPI_ISL_478050\|2020- | .................... | ...................A | .................... | .................... | .................... | .................... | 1 |
| D | 481 hCoV-19/Scotland/CVR2964/2020\|EPI_ISL_478051\|2020- | .................... | .................... | .................... | .................... | .................... | .................... | |
| D | 482 hCoV-19/Scotland/CVR3139/2020\|EPI_ISL_478053\|2020- | .................... | .................... | .................... | .................... | .................... | .................... | |
| D | 483 hCoV-19/Scotland/CVR3447/2020\|EPI_ISL_478058\|2020- | .................... | .................... | .................... | .................... | .................... | .................... | |
| D | 484 hCoV-19/Scotland/CVR3449/2020\|EPI_ISL_478059\|2020- | .................... | .................... | .................... | .................... | .................... | .................... | |
| D | 485 hCoV-19/Scotland/CVR3460/2020\|EPI_ISL_478065\|2020- | .................... | ...................A | .................... | .................... | .................... | .................... | 1 |
| D | 486 hCoV-19/Scotland/CVR3461/2020\|EPI_ISL_478066\|2020- | .................... | .................... | .................... | .................... | .................... | .................... | |
| D | 488 hCoV-19/Scotland/CVR3468/2020\|EPI_ISL_478071\|2020- | .................... | ...................A | .................... | .................... | .................... | .................... | 1 |
| D | 489 hCoV-19/Scotland/CVR3629/2020\|EPI_ISL_478079\|2020- | .................... | .................... | .................... | .................... | .................... | .................... | |
| D | 490 hCoV-19/Scotland/CVR3631/2020\|EPI_ISL_478080\|2020- | .................... | .................... | .................... | .................... | .................... | .................... | |
| D | 491 hCoV-19/Scotland/CVR3648/2020\|EPI_ISL_478084\|2020- | .................... | .................... | .................... | .................... | .................... | .................... | |
| D | 492 hCoV-19/Scotland/CVR3656/2020\|EPI_ISL_478085\|2020- | .................... | ...................A | .................... | .................... | .................... | .................... | 1 |
| D | 493 hCoV-19/Scotland/CVR3657/2020\|EPI_ISL_478086\|2020- | .................... | .................... | .................... | .................... | .................... | .................... | |
| D | 494 hCoV-19/Scotland/CVR3663/2020\|EPI_ISL_478089\|2020- | .................... | ...................A | .................... | .................... | .................... | .................... | 1 |
| D | 495 hCoV-19/Scotland/CVR3676/2020\|EPI_ISL_478090\|2020- | .................... | ...................A | .................... | .................... | .................... | .................... | 1 |
| D | 496 hCoV-19/Scotland/CVR3677/2020\|EPI_ISL_478091\|2020- | .................... | .................... | .................... | .................... | .................... | .................... | |
| D | 497 hCoV-19/Scotland/CVR3680/2020\|EPI_ISL_478093\|2020- | .................... | .................... | .................... | .................... | .................... | .................... | |
| D | 498 hCoV-19/Scotland/CVR3690/2020\|EPI_ISL_478094\|2020- | .................... | .................... | .................... | .................... | .................... | .................... | |

| | | | | | | | |
|---|---|---|---|---|---|---|---|
| D | 499 hCoV-19/Scotland/CVR3699/2020\|EPI_ISL_478095\|2020- | ···················· | ···················· | ···················· | ···················· | ···················· | ···················· |
| D | 500 hCoV-19/Scotland/CVR3706/2020\|EPI_ISL_478097\|2020- | ···················· | ···················· | ···················· | ···················· | ···················· | ···················· |
| D | 501 hCoV-19/Scotland/CVR3723/2020\|EPI_ISL_478098\|2020- | ···················· | ···················· | ···················· | ···················· | ···················· | ···················· |
| D | 502 hCoV-19/Wales/PHWC-164AB0/2020\|EPI_ISL_479344\|2020 | ···················· | ···················· | ···················· | ···················· | ···················· | ···················· |
| D | 503 hCoV-19/Wales/PHWC-164AED/2020\|EPI_ISL_479346\|2020 | ···················· | ···················· | ···················· | ···················· | ···················· | ···················· |
| D | 504 hCoV-19/Wales/PHWC-164B26/2020\|EPI_ISL_479349\|2020 | ···················· | ···················· | ···················· | ···················· | ···················· | ···················· |
| D | 505 hCoV-19/Wales/PHWC-164BAE/2020\|EPI_ISL_479355\|2020 | ···················· | ···················· | ···················· | ···················· | ···················· | ···················· |
| D | 506 hCoV-19/Wales/PHWC-164C23/2020\|EPI_ISL_479361\|2020 | ···················· | ···················· | ···················· | ···················· | ···················· | ···················· |
| D | 507 hCoV-19/Wales/PHWC-164C41/2020\|EPI_ISL_479362\|2020 | ···················· | ···················· | ···················· | ···················· | ···················· | ···················· |
| D | 508 hCoV-19/Wales/PHWC-164C7E/2020\|EPI_ISL_479365\|2020 | ···················· | ···················· | ···················· | ···················· | ···················· | ···················· |
| D | 510 hCoV-19/India/MH-NIV-15941/2020\|EPI_ISL_479504\|202 | ···················· | ···················· | ···················· | ···················· | ···················· | ···················· |
| D | 511 hCoV-19/India/MH-NIV-16503/2020\|EPI_ISL_479505\|202 | ···················· | ···················· | ···················· | ···················· | ···················· | ···················· |
| D | 513 hCoV-19/USA/MI-MDHHS-SC20591/2020\|EPI_ISL_462673\|2 | ···················· | ···················· | ···················· | ···················· | ···················· | ···················· |
| D | 514 hCoV-19/USA/MI-MDHHS-SC20592/2020\|EPI_ISL_462674\|2 | ···················· | ···················· | ···················· | ···················· | ···················· | ···················· |
| D | 515 hCoV-19/USA/MI-MDHHS-SC20593/2020\|EPI_ISL_462675\|2 | ···················· | ···················· | ···················· | ···················· | ···················· | ···················· |
| D | 516 hCoV-19/USA/MI-MDHHS-SC20596/2020\|EPI_ISL_462678\|2 | ···················· | ···················· | ···················· | ···················· | ···················· | ···················· |
| D | 517 hCoV-19/USA/MI-MDHHS-SC20597/2020\|EPI_ISL_462679\|2 | ···················· | ···················· | ···················· | ···················· | ···················· | ···················· |
| D | 518 hCoV-19/USA/MI-MDHHS-SC20598/2020\|EPI_ISL_462680\|2 | ···················· | ···················· | ···················· | ···················· | ···················· | ···················· |
| D | 519 hCoV-19/USA/MI-MDHHS-SC20599/2020\|EPI_ISL_462681\|2 | ···················· | ···················· | ···················· | ···················· | ···················· | ···················· |
| D | 520 hCoV-19/USA/MI-MDHHS-SC20601/2020\|EPI_ISL_462683\|2 | ···················· | ···················· | ···················· | ···················· | ···················· | ···················· |
| D | 521 hCoV-19/USA/MI-MDHHS-SC20602/2020\|EPI_ISL_462684\|2 | ···················· | ···················· | ···················· | ···················· | ···················· | ···················· |
| D | 522 hCoV-19/USA/MI-MDHHS-SC20603/2020\|EPI_ISL_462685\|2 | ···················· | ···················· | ···················· | ···················· | ···················· | ···················· |
| D | 523 hCoV-19/USA/MI-MDHHS-SC20604/2020\|EPI_ISL_462686\|2 | ···················· | ···················· | ···················· | ···················· | ···················· | ···················· |
| D | 524 hCoV-19/USA/MI-MDHHS-SC20605/2020\|EPI_ISL_462687\|2 | ···················· | ···················· | ···················· | ···················· | ···················· | ···················· |
| D | 525 hCoV-19/USA/MI-MDHHS-SC20606/2020\|EPI_ISL_462688\|2 | ···················· | ···················· | ···················· | ···················· | ···················· | ···················· |
| D | 526 hCoV-19/USA/MI-MDHHS-SC20607/2020\|EPI_ISL_462689\|2 | ···················· | ···················· | ···················· | ···················· | ···················· | ···················· |
| D | 527 hCoV-19/USA/MI-MDHHS-SC20608/2020\|EPI_ISL_462690\|2 | ···················· | ···················· | ···················· | ···················· | ···················· | ···················· |
| D | 528 hCoV-19/USA/MI-MDHHS-SC20609/2020\|EPI_ISL_462691\|2 | ···················· | ···················· | ···················· | ···················· | ···················· | ···················· |
| D | 529 hCoV-19/USA/MI-MDHHS-SC20610/2020\|EPI_ISL_462692\|2 | ···················· | ···················· | ···················· | ···················· | ···················· | ···················· |
| D | 530 hCoV-19/USA/MI-MDHHS-SC20611/2020\|EPI_ISL_462693\|2 | ···················· | ···················· | ···················· | ···················· | ···················· | ···················· |
| D | 531 hCoV-19/USA/MI-MDHHS-SC20612/2020\|EPI_ISL_462694\|2 | ···················· | ···················· | ···················· | ···················· | ···················· | ···················· |
| D | 532 hCoV-19/USA/MI-MDHHS-SC20613/2020\|EPI_ISL_462695\|2 | ···················· | ···················· | ···················· | ···················· | ···················· | ···················· |
| D | 533 hCoV-19/USA/MI-MDHHS-SC20614/2020\|EPI_ISL_462696\|2 | ···················· | ···················· | ···················· | ···················· | ···················· | ···················· |
| D | 534 hCoV-19/USA/MI-MDHHS-SC20615/2020\|EPI_ISL_462697\|2 | ···················· | ···················· | ···················· | ···················· | ···················· | ···················· |
| D | 535 hCoV-19/USA/MI-MDHHS-SC20616/2020\|EPI_ISL_462698\|2 | ···················· | ···················· | ···················· | ···················· | ···················· | ···················· |
| D | 536 hCoV-19/USA/MI-MDHHS-SC20617/2020\|EPI_ISL_462699\|2 | ···················· | ···················· | ···················· | ···················· | ···················· | ···················· |
| D | 537 hCoV-19/England/20149045004/2020\|EPI_ISL_466200\|20 | ···················· | ···················· | ···················· | ···················· | ···················· | ···················· |
| D | 538 hCoV-19/England/20149048604/2020\|EPI_ISL_466201\|20 | ···················· | ···················· | ···················· | ···················· | ···················· | NN·················· |
| D | 539 hCoV-19/England/20149049104/2020\|EPI_ISL_466203\|20 | ···················· | ···················· | ···················· | ···················· | ···················· | ········N····N······ |
| D | 541 hCoV-19/England/20149051704/2020\|EPI_ISL_466205\|20 | ···················· | ···················· | ···················· | ···················· | ···················· | ···················· |
| D | 542 hCoV-19/England/20149052404/2020\|EPI_ISL_466206\|20 | ···················· | ···················· | ···················· | ···················· | ···················· | ···················· |
| D | 543 hCoV-19/England/20149052604/2020\|EPI_ISL_466207\|20 | ···················· | ···················· | ···················· | ···················· | ···················· | ···················· |
| D | 544 hCoV-19/England/20149052904/2020\|EPI_ISL_466208\|20 | ···················· | ···················· | ···················· | ···················· | ···················· | ···················· |
| D | 545 hCoV-19/England/20149054004/2020\|EPI_ISL_466212\|20 | ···················· | ···················· | ···················· | ···················· | ···················· | ···················· |
| D | 546 hCoV-19/England/20149054104/2020\|EPI_ISL_466213\|20 | ···················· | ···················· | ···················· | ···················· | ···················· | ···················· |
| D | 547 hCoV-19/England/20149054404/2020\|EPI_ISL_466216\|20 | ···················· | ···················· | ···················· | ···················· | ···················· | ···················· |
| D | 548 hCoV-19/England/20149054504/2020\|EPI_ISL_466217\|20 | ···················· | ···················· | ···················· | ···················· | ···················· | ···················· |
| D | 550 hCoV-19/England/20149055204/2020\|EPI_ISL_466219\|20 | ···················· | ···················· | ···················· | ···················· | ···················· | ···················· |
| D | 551 hCoV-19/England/20150032604/2020\|EPI_ISL_466224\|20 | ···T················ | ···················· | ···················· | ···················· | ···················· | ···················· |
| D | 554 hCoV-19/England/20150039804/2020\|EPI_ISL_466228\|20 | ···················· | ···················· | ···················· | ···················· | ·····C·············· | ···R················ |
| D | 556 hCoV-19/England/20150041004/2020\|EPI_ISL_466231\|20 | ···················· | ···················· | ···················· | ···················· | ···················· | ···················· |
| D | 557 hCoV-19/England/20152000202/2020\|EPI_ISL_466232\|20 | ···················· | ···················· | ···················· | ···················· | ··NN················ | ···················· |
| D | 558 hCoV-19/England/20152046204/2020\|EPI_ISL_466233\|20 | ···················· | ···················· | ···················· | ···················· | ···················· | ···················· |
| D | 559 hCoV-19/England/20152047704/2020\|EPI_ISL_466234\|20 | ···················· | ···················· | ···················· | ···················· | ···················· | ···················· |
| D | 560 hCoV-19/England/20152054104/2020\|EPI_ISL_466235\|20 | ···················· | ···················· | ···················· | ···················· | ···················· | ···················· |
| D | 561 hCoV-19/South Africa/KRISP-0150/2020\|EPI_ISL_46750 | ···················· | ···················· | ···················· | ···················· | ···················· | ···················· |
| D | 562 hCoV-19/South Africa/KRISP-0151/2020\|EPI_ISL_46750 | ···················· | ···················· | ···················· | ···················· | ···················· | ···················· |
| D | 563 hCoV-19/South Africa/KRISP-0152/2020\|EPI_ISL_46750 | ···················· | ···················· | ···················· | ···················· | ···················· | ···················· |
| D | 564 hCoV-19/South Africa/KRISP-0153/2020\|EPI_ISL_46750 | ···················· | ···················· | ···················· | ···················· | ···················· | ···················· |
| D | 565 hCoV-19/South Africa/KRISP-0154/2020\|EPI_ISL_46750 | ···················· | ···················· | ···················· | ···················· | ···················· | ···················· |
| D | 566 hCoV-19/South Africa/KRISP-0156/2020\|EPI_ISL_46750 | ···················· | ···················· | ···················· | ···················· | ···················· | ···················· |
| D | 567 hCoV-19/South Africa/KRISP-0157/2020\|EPI_ISL_46750 | ···················· | ···················· | ···················· | ···················· | ···················· | ···················· |
| D | 568 hCoV-19/South Africa/KRISP-0158/2020\|EPI_ISL_46750 | ···················· | ···················· | ···················· | ···················· | ···················· | ···················· |
| D | 569 hCoV-19/South Africa/KRISP-0159/2020\|EPI_ISL_46750 | ···················· | ···················· | ···················· | ···················· | ···················· | ···················· |
| D | 570 hCoV-19/South Africa/KRISP-0161/2020\|EPI_ISL_46751 | ···················· | ···················· | ···················· | ···················· | ···················· | ···················· |
| D | 571 hCoV-19/South Africa/KRISP-0162/2020\|EPI_ISL_46751 | ···················· | ···················· | ···················· | ···················· | ···················· | ···················· |
| D | 572 hCoV-19/South Africa/KRISP-0163/2020\|EPI_ISL_46751 | ···················· | ···················· | ···················· | ···················· | ···················· | ···················· |
| D | 573 hCoV-19/South Africa/KRISP-0164/2020\|EPI_ISL_46751 | ···················· | ···················· | ···················· | ···················· | ···················· | ···················· |
| D | 574 hCoV-19/South Africa/KRISP-0165/2020\|EPI_ISL_46751 | ···················· | ···················· | ···················· | ···················· | ···················· | ···················· |
| D | 575 hCoV-19/South Africa/KRISP-0166/2020\|EPI_ISL_46751 | ···················· | ···················· | ···················· | ···················· | ···················· | ···················· |
| D | 576 hCoV-19/South Africa/KRISP-0167/2020\|EPI_ISL_46751 | ···················· | ···················· | ···················· | ···················· | ···················· | ···················· |
| D | 577 hCoV-19/South Africa/KRISP-0168/2020\|EPI_ISL_46751 | ···················· | ···················· | ···················· | ···················· | ···················· | ···················· |
| D | 578 hCoV-19/South Africa/KRISP-0169/2020\|EPI_ISL_46751 | ···················· | ···················· | ···················· | ···················· | ···················· | ···················· |
| D | 579 hCoV-19/South Africa/KRISP-0174/2020\|EPI_ISL_46752 | ···················· | ···················· | ···················· | ···················· | ···················· | ···················· |
| D | 580 hCoV-19/USA/NM-UNM-00064/2020\|EPI_ISL_467529\|2020- | ···················· | ···················· | ···················· | ···················· | ···················· | ···················· |
| D | 581 hCoV-19/USA/NM-UNM-00079/2020\|EPI_ISL_467536\|2020- | ···················· | ···················· | ···················· | ···················· | ···················· | ···················· |
| D | 582 hCoV-19/USA/NM-UNM-00100/2020\|EPI_ISL_467545\|2020- | ···················· | ···················· | ···················· | ···················· | ···················· | ···················· |
| D | 583 hCoV-19/USA/NM-UNM-00112/2020\|EPI_ISL_467551\|2020- | ···················· | ···················· | ···················· | ···················· | ···················· | ···················· |

| | | | | | | | | | |
|---|---|---|---|---|---|---|---|---|---|
| D | 584 hCoV-19/USA/NM-UNM-00113/2020\|EPI_ISL_467552\|2020- | .................... | .................... | .................... | .................... | .................... | .................... | | |
| D | 585 hCoV-19/USA/NM-UNM-00136/2020\|EPI_ISL_467563\|2020- | .................... | .................... | .................... | .................... | .................... | .................... | | |
| D | 586 hCoV-19/USA/NM-UNM-00137/2020\|EPI_ISL_467564\|2020- | .................... | .................... | .................... | .................... | .................... | .................... | | |
| D | 598 hCoV-19/Spain/AR-IBV-2422/2020\|EPI_ISL_468815\|2020 | .................... | .................... | .................... | .................... | NNN................. | .................... | | 1 |
| D | 612 hCoV-19/Spain/AR-IBV-2453/2020\|EPI_ISL_468831\|2020 | .................... | .................... | .................... | .................... | .................... | .................... | | |
| D | 619 hCoV-19/Spain/AR-IBV-2387/2020\|EPI_ISL_468840\|2020 | .................... | .................... | .................... | .................... | .................... | .................... | | |
| D | 630 hCoV-19/Spain/AR-IBV-2423/2020\|EPI_ISL_468859\|2020 | .................... | .................... | .................... | .................... | .................... | .................... | | |
| D | 631 hCoV-19/Spain/PV-IBV-2151/2020\|EPI_ISL_468860\|2020 | .................... | .................... | .................... | .................... | .................... | .................... | | |
| D | 643 hCoV-19/Spain/PV-IBV-2108/2020\|EPI_ISL_468873\|2020 | .................... | .................... | .................... | .................... | .................... | .................... | | |
| D | 651 hCoV-19/Spain/PV-IBV-2113/2020\|EPI_ISL_468881\|2020 | .................... | .................... | .................... | .................... | .................... | .................... | | |
| D | 654 hCoV-19/Spain/PV-IBV-2137/2020\|EPI_ISL_468885\|2020 | .................... | .................... | .................... | .................... | .................... | .................... | | |
| D | 658 hCoV-19/Spain/PV-IBV-2121/2020\|EPI_ISL_468891\|2020 | .................... | .................... | .................... | .................... | .................... | .................... | | |
| D | 659 hCoV-19/Spain/PV-IBV-2114/2020\|EPI_ISL_468892\|2020 | .................... | .................... | .................... | .................... | .................... | .................... | | |
| D | 664 hCoV-19/Spain/PV-IBV-2118/2020\|EPI_ISL_468897\|2020 | .................... | .................... | .................... | .................... | .................... | .................... | | |
| D | 666 hCoV-19/England/LIVE-9F6B9/2020\|EPI_ISL_472000\|202 | .................... | .................... | .................... | .................... | .................... | .................... | | |
| D | 667 hCoV-19/England/LIVE-9FCE4/2020\|EPI_ISL_472057\|202 | .................... | .................... | .................... | .................... | .................... | .................... | | |
| D | 668 hCoV-19/England/LIVE-A00F3/2020\|EPI_ISL_472090\|202 | .................... | .A.................. | .................... | .................... | .................... | .................... | | |
| D | 669 hCoV-19/England/LIVE-A015A/2020\|EPI_ISL_472091\|202 | .................... | .................... | .................... | .................... | .................... | .................... | | |
| D | 670 hCoV-19/England/LIVE-A01A5/2020\|EPI_ISL_472092\|202 | .................... | .................... | .................... | .................... | .................... | .................... | | |
| D | 671 hCoV-19/England/LIVE-A01B4/2020\|EPI_ISL_472093\|202 | .................... | .A.................. | .................... | .................... | .................... | .................... | | |
| D | 672 hCoV-19/England/LIVE-A01E1/2020\|EPI_ISL_472096\|202 | .................... | .................... | .................... | .................... | .................... | .................... | | |
| D | 673 hCoV-19/England/LIVE-A0248/2020\|EPI_ISL_472097\|202 | .................... | .................... | .................... | .................... | .................... | .................... | | |
| D | 674 hCoV-19/England/LIVE-A0293/2020\|EPI_ISL_472098\|202 | .................... | .................... | .................... | .................... | .................... | .................... | | |
| D | 675 hCoV-19/England/LIVE-A02C0/2020\|EPI_ISL_472099\|202 | .................... | .................... | .................... | .................... | .................... | .................... | | |
| D | 676 hCoV-19/England/BIRM-5F6D3/2020\|EPI_ISL_473321\|202 | .................... | .................... | .................... | .................... | .................... | .................... | | |
| D | 677 hCoV-19/England/BIRM-5F70D/2020\|EPI_ISL_473322\|202 | .................... | .................... | .................... | .................... | .................... | .................... | | |
| D | 678 hCoV-19/England/BIRM-5F71C/2020\|EPI_ISL_473323\|202 | .................... | .................... | .................... | .................... | .................... | .................... | | |
| D | 679 hCoV-19/England/BIRM-5F73A/2020\|EPI_ISL_473324\|202 | .................... | .................... | .................... | .................... | .................... | .................... | | |
| D | 680 hCoV-19/England/BIRM-5F785/2020\|EPI_ISL_473328\|202 | .................... | .................... | .................... | .................... | .................... | .................... | | |
| D | 681 hCoV-19/England/BIRM-5F794/2020\|EPI_ISL_473329\|202 | .................... | .................... | .................... | .................... | .................... | .................... | | |
| D | 682 hCoV-19/England/BIRM-5F7A3/2020\|EPI_ISL_473330\|202 | .................... | .................... | .................... | .................... | .................... | .................... | | |
| D | 683 hCoV-19/England/BIRM-5F7B2/2020\|EPI_ISL_473331\|202 | .................... | .................... | .................... | .................... | .................... | .................... | | |
| D | 684 hCoV-19/England/BIRM-5FA40/2020\|EPI_ISL_473365\|202 | .................... | .................... | .................... | .................... | .................... | .................... | | |
| D | 685 hCoV-19/England/BIRM-5FAC8/2020\|EPI_ISL_473371\|202 | .................... | .................... | .................... | .................... | .................... | .................... | | |
| D | 686 hCoV-19/England/BIRM-5FAF5/2020\|EPI_ISL_473374\|202 | .................... | .................... | .................... | .................... | .................... | .................... | | |
| D | 687 hCoV-19/England/BIRM-5FB2F/2020\|EPI_ISL_473376\|202 | .................... | .................... | .................... | .................... | .................... | .................... | | |
| D | 689 hCoV-19/England/BIRM-5FB5C/2020\|EPI_ISL_473379\|202 | .................... | .................... | .................... | .................... | .................... | .................... | | |
| D | 690 hCoV-19/England/BIRM-5FB7A/2020\|EPI_ISL_473381\|202 | .................... | .................... | .................... | .................... | .................... | .................... | | |
| D | 691 hCoV-19/England/BIRM-5FB89/2020\|EPI_ISL_473382\|202 | .................... | .................... | .................... | .................... | .................... | .................... | | |
| D | 692 hCoV-19/England/BIRM-5FB98/2020\|EPI_ISL_473383\|202 | .................... | .................... | .................... | .................... | .................... | .................... | | |
| D | 694 hCoV-19/England/BIRM-5FBB6/2020\|EPI_ISL_473385\|202 | .................... | .................... | .................... | .................... | .................... | .................... | | |
| D | 695 hCoV-19/England/BIRM-5FBC5/2020\|EPI_ISL_473386\|202 | .................... | .................... | .................... | .................... | .................... | .................... | | |
| D | 696 hCoV-19/England/BIRM-5FBE3/2020\|EPI_ISL_473388\|202 | .................... | .................... | .................... | .................... | .................... | .................... | | |
| D | 697 hCoV-19/England/BIRM-5FBF2/2020\|EPI_ISL_473389\|202 | .................... | .................... | .................... | .................... | .................... | .................... | | |
| D | 698 hCoV-19/England/BIRM-5FC0E/2020\|EPI_ISL_473390\|202 | .................... | .................... | .................... | .................... | .................... | .................... | | |
| D | 699 hCoV-19/England/BIRM-5FC59/2020\|EPI_ISL_473392\|202 | .................... | .................... | .................... | .................... | .................... | .................... | | |
| D | 700 hCoV-19/England/BIRM-5FCA4/2020\|EPI_ISL_473394\|202 | .................... | .................... | .................... | .................... | .................... | .................... | | |
| D | 701 hCoV-19/Wales/PHWC-36362/2020\|EPI_ISL_474656\|2020- | .................... | .................... | .................... | .................... | .................... | .................... | | |
| D | 702 hCoV-19/Wales/PHWC-36371/2020\|EPI_ISL_474657\|2020- | .................... | .................... | .................... | .................... | .................... | .................... | | |
| D | 703 hCoV-19/Wales/PHWC-36380/2020\|EPI_ISL_474658\|2020- | .................... | .................... | .................... | .................... | .................... | .................... | | |
| D | 704 hCoV-19/Wales/PHWC-363AE/2020\|EPI_ISL_474660\|2020- | .................... | .................... | .................... | .................... | .................... | .................... | | |
| D | 705 hCoV-19/Wales/PHWC-366B4/2020\|EPI_ISL_474699\|2020- | .................... | .................... | .................... | .................... | .................... | .................... | | |
| D | 706 hCoV-19/Singapore/452/2020\|EPI_ISL_475983\|2020-03- | .................... | .................... | .................... | .................... | .................... | .................... | | |
| D | 708 hCoV-19/Singapore/455/2020\|EPI_ISL_475986\|2020-03- | .................... | .................... | .................... | .................... | .................... | .................... | | |
| D | 709 hCoV-19/Singapore/459/2020\|EPI_ISL_475990\|2020-04- | .................... | .................... | .................... | .................... | .................... | .................... | | |
| D | 710 hCoV-19/Singapore/460/2020\|EPI_ISL_475991\|2020-04- | .................... | .................... | .................... | .................... | .................... | .................... | | |
| D | 711 hCoV-19/Singapore/462/2020\|EPI_ISL_475993\|2020-04- | .................... | .................... | .................... | .................... | .................... | .................... | | |
| D | 712 hCoV-19/Scotland/EDB6062/2020\|EPI_ISL_478200\|2020- | .................... | ...................A | .................... | .................... | .................... | .................... | 1 | |
| D | 713 hCoV-19/Scotland/EDB6064/2020\|EPI_ISL_478201\|2020- | .................... | ...................A | .................... | .................... | .................... | .................... | 1 | |
| D | 714 hCoV-19/Scotland/EDB6066/2020\|EPI_ISL_478203\|2020- | .................... | ...................A | .................... | .................... | .................... | .................... | 1 | |
| D | 715 hCoV-19/Scotland/EDB6068/2020\|EPI_ISL_478204\|2020- | .................... | .................... | .................... | .................... | .................... | .................... | | |
| D | 716 hCoV-19/Scotland/EDB6070/2020\|EPI_ISL_478205\|2020- | .................... | ...................A | .................... | .................... | .................... | .................... | 1 | |
| D | 717 hCoV-19/Scotland/EDB6078/2020\|EPI_ISL_478206\|2020- | .................... | ...................A | .................... | .................... | .................... | .................... | 1 | |
| D | 718 hCoV-19/Scotland/EDB6081/2020\|EPI_ISL_478207\|2020- | .................... | ...................A | .................... | .................... | .................... | .................... | 1 | |
| D | 719 hCoV-19/Scotland/EDB6083/2020\|EPI_ISL_478208\|2020- | .................... | .A.................. | .................... | .................... | .................... | .................... | | |
| D | 720 hCoV-19/Scotland/EDB6085/2020\|EPI_ISL_478209\|2020- | .................... | ...................A | .................... | .................... | .................... | .................... | 1 | |
| D | 721 hCoV-19/Scotland/EDB6086/2020\|EPI_ISL_478210\|2020- | .................... | .A.................. | .................... | .................... | .................... | .................... | | |
| D | 722 hCoV-19/Scotland/EDB6088/2020\|EPI_ISL_478211\|2020- | .................... | ...................A | .................... | .................... | .................... | .................... | 1 | |
| D | 723 hCoV-19/Scotland/EDB6089/2020\|EPI_ISL_478212\|2020- | .................... | .................... | .................... | .................... | .................... | .................... | | |
| D | 724 hCoV-19/Scotland/EDB6090/2020\|EPI_ISL_478213\|2020- | .................... | ...................A | .................... | .................... | .................... | .................... | 1 | |
| D | 725 hCoV-19/Scotland/EDB6092/2020\|EPI_ISL_478214\|2020- | .................... | .................... | .................... | .................... | .................... | .................... | | |
| D | 726 hCoV-19/Scotland/EDB6093/2020\|EPI_ISL_478215\|2020- | .................... | .A.................. | .................... | .................... | .................... | .................... | | |
| D | 727 hCoV-19/India/MH-NIV-15115/2020\|EPI_ISL_479500\|202 | .................... | .................... | .................... | .................... | .................... | .................... | | |
| D | 728 hCoV-19/India/MH-NIV-15351/2020\|EPI_ISL_479501\|202 | .................... | .................... | .................... | .................... | .................... | .................... | | |
| D | 729 hCoV-19/India/MH-NIV-15768/2020\|EPI_ISL_479502\|202 | .................... | .................... | .................... | .................... | .................... | .................... | | |
| D | 730 hCoV-19/India/MH-NIV-17002/2020\|EPI_ISL_479507\|202 | .................... | .................... | .................... | .................... | .................... | .................... | | |
| D | 731 hCoV-19/India/MH-NIV-17547/2020\|EPI_ISL_479508\|202 | .................... | .................... | .................... | .................... | .................... | .................... | | |
| D | 732 hCoV-19/India/MH-NIV-17570/2020\|EPI_ISL_479509\|202 | .................... | .................... | .................... | .................... | .................... | .................... | | |

```
D 734 hCoV-19/India/MH-NIV-18728/2020|EPI_ISL_479511|202 .................... .................... ..................... .................... .................... ....................
D 735 hCoV-19/India/MH-NIV-19746/2020|EPI_ISL_479512|202 .................... .................... ..................... .................... .................... ....................
D 736 hCoV-19/India/MH-NIV-20134/2020|EPI_ISL_479513|202 .................... .................... ..................... .................... .................... ....................
D 737 hCoV-19/India/MH-NIV-20608/2020|EPI_ISL_479514|202 .................... .................... ..................... .................... .................... ....................
D 740 hCoV-19/India/MH-NIV-21455/2020|EPI_ISL_479517|202 .................... .................... ..................... .................... .................... ....................
D 741 hCoV-19/India/MH-NIV-22048/2020|EPI_ISL_479518|202 .................... .................... ..................... .................... .................... ....................
D 742 hCoV-19/India/MH-NIV-22552/2020|EPI_ISL_479519|202 .................... .................... ..................... .................... .................... ....................
D 743 hCoV-19/India/MH-NIV-22876/2020|EPI_ISL_479520|202 .................... .................... ..................... .................... .................... ....................
D 744 hCoV-19/India/MH-NIV-23098/2020|EPI_ISL_479521|202 .................... .................... ..................... .................... .................... ....................
D 745 hCoV-19/India/MH-NIV-23721/2020|EPI_ISL_479522|202 .................... .................... ..................... .................... .................... ....................
D 746 hCoV-19/India/MH-NIV-24174/2020|EPI_ISL_479523|202 .................... .................... ..................... .................... .................... ....................
D 749 hCoV-19/India/MH-NIV-28147/2020|EPI_ISL_479527|202 .................... .................... ..................... .................... .................... ....................
D 751 hCoV-19/India/MH-NIV-28815/2020|EPI_ISL_479529|202 .................... .................... ..................... .................... .................... ....................
D 753 hCoV-19/India/MH-NIV-29053/2020|EPI_ISL_479531|202 .................... .................... ..................... .................... .................... ....................
D 755 hCoV-19/India/MH-NIV-31191/2020|EPI_ISL_479533|202 .................... .................... ..................... .................... .................... ....................
D 758 hCoV-19/India/MH-NIV-36444/2020|EPI_ISL_479536|202 .................... .................... ..................... .................... .................... ....................
D 759 hCoV-19/India/MH-NIV-36461/2020|EPI_ISL_479537|202 .................... .................... ..................... .................... .................... ....................
D 760 hCoV-19/India/MH-NIV-37486/2020|EPI_ISL_479538|202 .................... .................... ..................... .................... .................... ....................
D 761 hCoV-19/India/MH-NIV-37502/2020|EPI_ISL_479539|202 .................... .................... ..................... .................... .................... ....................
D 762 hCoV-19/India/MH-NIV-37681/2020|EPI_ISL_479540|202 .................... .................... ..................... .................... .................... ....................
D 763 hCoV-19/India/MH-NIV-37985/2020|EPI_ISL_479541|202 .................... .................... ..................... .................... .................... ....................
D 765 hCoV-19/India/MH-NIV-40837/2020|EPI_ISL_479543|202 .................... .................... ..................... .................... .................... ....................
D 766 hCoV-19/India/MH-NIV-42694/2020|EPI_ISL_479544|202 .................... .................... ..................... .................... .................... ....................
D 767 hCoV-19/India/MH-NIV-46430/2020|EPI_ISL_479545|202 .................... .................... ..................... .................... .................... ....................
D 768 hCoV-19/India/MH-NIV-44241/2020|EPI_ISL_479546|202 .................... .................... ..................... .................... .................... ....................
D 770 hCoV-19/Canada/QC_AO7/2020|EPI_ISL_463922|2020-04- .................... .................... ..................... .................... .................... ....................
D 771 hCoV-19/Canada/QC_AO9/2020|EPI_ISL_463923|2020-04- .................... .................... ..................... .................... .................... ....................
D 772 hCoV-19/Canada/QC_AQ1/2020|EPI_ISL_463927|2020-04- .................... .................... ..................... .................... .................... ....................
D 774 hCoV-19/Canada/QC_AQ5/2020|EPI_ISL_463929|2020-04- .................... .................... ..................... .................... .................... ....................
D 775 hCoV-19/Canada/QC_AQ6/2020|EPI_ISL_463930|2020-04- .................... .................... ..................... .................... .................... ....................
D 776 hCoV-19/Canada/QC_AR3/2020|EPI_ISL_463931|2020-04- .................... .................... ..................... .................... .................... ....................
D 777 hCoV-19/Canada/QC_AR6/2020|EPI_ISL_463933|2020-04- .................... .................... ..................... .................... .................... ....................
D 779 hCoV-19/Canada/QC_AS4/2020|EPI_ISL_463936|2020-04- .................... .................... ..................... .................... .................... ....................
D 780 hCoV-19/Canada/QC_AT3/2020|EPI_ISL_463940|2020-03- .................... .................... ..................... .................... .................... ....................
D 782 hCoV-19/Canada/QC_AT7/2020|EPI_ISL_463942|2020-04- .................... .................... ..................... .................... .................... ....................
D 783 hCoV-19/Canada/QC_AT8/2020|EPI_ISL_463943|2020-04- .................... .................... ..................... .................... .................... ....................
D 784 hCoV-19/Canada/QC_AU7/2020|EPI_ISL_463946|2020-04- .................... .................... ..................... .................... .................... ....................
D 785 hCoV-19/Canada/QC_AU8/2020|EPI_ISL_463947|2020-04- .................... .................... ..................... .................... .................... ....................
D 786 hCoV-19/Canada/QC_AU9/2020|EPI_ISL_463948|2020-04- .................... .................... ..................... .................... .................... ....................
D 787 hCoV-19/Canada/QC_AX6/2020|EPI_ISL_463953|2020-04- .................... .................... ..................... .................... .................... ....................
D 788 hCoV-19/Canada/QC_AY1/2020|EPI_ISL_463954|2020-04- .................... .................... ..................... .................... .................... ....................
D 789 hCoV-19/Canada/QC_AY8/2020|EPI_ISL_463956|2020-04- .................... .................... ..................... .................... .................... ....................
D 790 hCoV-19/Canada/QC_AY9/2020|EPI_ISL_463957|2020-04- .................... .................... ..................... .................... .................... ....................
D 791 hCoV-19/Canada/QC_AZ9/2020|EPI_ISL_463961|2020-04- .................... .................... ..................... .................... .................... ....................
D 792 hCoV-19/Canada/QC_BA5/2020|EPI_ISL_463962|2020-04- .................... .................... ..................... .................... .................... ....................
D 793 hCoV-19/Canada/QC_BA6/2020|EPI_ISL_463963|2020-04- .................... .................... ..................... .................... .................... ....................
D 794 hCoV-19/Canada/QC_BB7/2020|EPI_ISL_463965|2020-04- .................... .................... ..................... .................... .................... ....................
D 795 hCoV-19/Canada/QC_BC5/2020|EPI_ISL_463968|2020-04- .................... .................... ..................... .................... .................... ....................
D 796 hCoV-19/Canada/ON_MU-S1/2020|EPI_ISL_463970|2020-0 .................... .................... ..................... .................... .................... ....................
D 797 hCoV-19/Canada/ON_MU-S13/2020|EPI_ISL_463971|2020- .................... .................... ..................... .................... .................... ....................
D 798 hCoV-19/Canada/ON_MU-S26/2020|EPI_ISL_463975|2020- .................... .................... ..................... .................... .................... ....................
D 799 hCoV-19/Canada/ON_MU-S284/2020|EPI_ISL_463976|2020 .................... .................... ..................... .................... .................... ....................
D 800 hCoV-19/Canada/ON_MU-S29/2020|EPI_ISL_463977|2020- .................... .................... ..................... .................... .................... ....................
D 801 hCoV-19/Canada/ON_MU-S306/2020|EPI_ISL_463979|2020 .................... .................... ..................... .................... .................... ....................
D 802 hCoV-19/Canada/ON_MU-S312/2020|EPI_ISL_463980|2020 .................... .................... ..................... .................... .................... ....................
D 803 hCoV-19/Canada/ON_MU-S328/2020|EPI_ISL_463981|2020 .................... .................... ..................... .................... .................... ....................
D 804 hCoV-19/Canada/ON_MU-S349/2020|EPI_ISL_463982|2020 .................... .................... ..................... .................... .................... ....................
D 805 hCoV-19/Canada/ON_MU-S40/2020|EPI_ISL_463984|2020- .................... .................... ..................... .................... .................... ....................
D 806 hCoV-19/Canada/ON_MU-S5/2020|EPI_ISL_463988|2020-0 .................... .................... ..................... .................... .................... ....................
D 807 hCoV-19/Canada/ON_MU-S51/2020|EPI_ISL_463989|2020- .................... .................... ..................... .................... .................... ....................
D 808 hCoV-19/Canada/ON_MU-S63/2020|EPI_ISL_463991|2020- .................... .................... ..................... .................... .................... ....................
D 809 hCoV-19/Canada/ON_MU-S679/2020|EPI_ISL_463992|2020 .................... .................... ..................... .................... .................... ....................
D 810 hCoV-19/Canada/ON_MU-S7/2020|EPI_ISL_463993|2020-0 .................... .................... ..................... .................... .................... ....................
D 811 hCoV-19/Canada/ON_MU-S82/2020|EPI_ISL_463994|2020- .................... .................... ..................... .................... .................... ....................
D 812 hCoV-19/Canada/ON-UHTC_0078/2020|EPI_ISL_463995|20 .................... .................... ..................... .................... .................... ....................
D 813 hCoV-19/Canada/ON-UHTC_0071/2020|EPI_ISL_463996|20 .................... .................... ..................... .................... .................... ....................
D 814 hCoV-19/Canada/ON-UHTC_0067/2020|EPI_ISL_463997|20 .................... .................... ..................... .................... .................... ....................
D 815 hCoV-19/Canada/ON-UHTC_0082/2020|EPI_ISL_463998|20 .................... .................... ..................... .................... .................... ....................
D 816 hCoV-19/Canada/ON-UHTC_0075/2020|EPI_ISL_463999|20 .................... .................... ..................... .................... .................... ....................
D 817 hCoV-19/England/20149008904/2020|EPI_ISL_466141|20 .................... .................... ..................... .................... .................... ....................
D 818 hCoV-19/England/20149009004/2020|EPI_ISL_466142|20 .................... .................... ..................... .................... .................... ....................
D 819 hCoV-19/England/20149009204/2020|EPI_ISL_466144|20 .................... .................... ..................... .................... .................... ....................
D 820 hCoV-19/England/20149009304/2020|EPI_ISL_466145|20 .................... .................... ..................... .................... .................... ....................
D 821 hCoV-19/England/20149009404/2020|EPI_ISL_466146|20 .................... .................... ..................... .................... .................... ....................
D 824 hCoV-19/England/20149009704/2020|EPI_ISL_466149|20 .................... .................... ..................... .................... .................... ....................
D 825 hCoV-19/England/20149011904/2020|EPI_ISL_466151|20 .................... .................... ..................... .................... .................... ....................
D 826 hCoV-19/England/20149012004/2020|EPI_ISL_466152|20 .................... .................... ..................... .................... .................... ....................
D 827 hCoV-19/England/20149012504/2020|EPI_ISL_466153|20 .................... .................... ..................... .................... .................... ....................
```

| | | | | | | | |
|---|---|---|---|---|---|---|---|
| D | 829 hCoV-19/England/20149013504/2020\|EPI_ISL_466156\|20 | ···················· | ···················· | ···················· | ···················· | ···················· | ···················· |
| D | 830 hCoV-19/England/20149013604/2020\|EPI_ISL_466157\|20 | ···················· | ···················· | ···················· | ···················· | ···················· | ···················· |
| D | 831 hCoV-19/England/20149013704/2020\|EPI_ISL_466158\|20 | ···················· | ···················· | ···················· | ···················· | ···················· | ···················· |
| D | 832 hCoV-19/England/20149014504/2020\|EPI_ISL_466159\|20 | ···················· | ···················· | ···················· | ···················· | ···················· | ···················· |
| D | 833 hCoV-19/South Africa/KRISP-0118/2020\|EPI_ISL_46747 | ···················· | ···················· | ···················· | ···················· | ···················· | ···················· |
| D | 834 hCoV-19/South Africa/KRISP-0119/2020\|EPI_ISL_46747 | ···················· | ···················· | ···················· | ···················· | ···················· | ···················· |
| D | 835 hCoV-19/South Africa/KRISP-0120/2020\|EPI_ISL_46747 | ···················· | ···················· | ···················· | ···················· | ···················· | ···················· |
| D | 836 hCoV-19/South Africa/KRISP-0121/2020\|EPI_ISL_46747 | ···················· | ···················· | ···················· | ···················· | ···················· | ···················· |
| D | 837 hCoV-19/South Africa/KRISP-0122/2020\|EPI_ISL_46747 | ···················· | ···················· | ···················· | ···················· | ···················· | ···················· |
| D | 838 hCoV-19/South Africa/KRISP-0123/2020\|EPI_ISL_46747 | ···················· | ···················· | ···················· | ···················· | ···················· | ···················· |
| D | 839 hCoV-19/South Africa/KRISP-0124/2020\|EPI_ISL_46747 | ···················· | ···················· | ···················· | ···················· | ···················· | ···················· |
| D | 840 hCoV-19/South Africa/KRISP-0125/2020\|EPI_ISL_46747 | ···················· | ···················· | ···················· | ···················· | ···················· | ···················· |
| D | 841 hCoV-19/South Africa/KRISP-0126/2020\|EPI_ISL_46747 | ···················· | ···················· | ···················· | ···················· | ···················· | ···················· |
| D | 842 hCoV-19/South Africa/KRISP-0127/2020\|EPI_ISL_46747 | ···················· | ···················· | ···················· | ···················· | ···················· | ···················· |
| D | 843 hCoV-19/South Africa/KRISP-0128/2020\|EPI_ISL_46748 | ···················· | ···················· | ···················· | ···················· | ···················· | ···················· |
| D | 844 hCoV-19/South Africa/KRISP-0129/2020\|EPI_ISL_46748 | ···················· | ···················· | ···················· | ···················· | ···················· | ···················· |
| D | 845 hCoV-19/South Africa/KRISP-0130/2020\|EPI_ISL_46748 | ···················· | ···················· | ···················· | ···················· | ···················· | ···················· |
| D | 846 hCoV-19/South Africa/KRISP-0131/2020\|EPI_ISL_46748 | ···················· | ···················· | ···················· | ···················· | ···················· | ···················· |
| D | 847 hCoV-19/South Africa/KRISP-0132/2020\|EPI_ISL_46748 | ···················· | ···················· | ···················· | ···················· | ···················· | ···················· |
| D | 848 hCoV-19/South Africa/KRISP-0133/2020\|EPI_ISL_46748 | ···················· | ···················· | ···················· | ···················· | ···················· | ···················· |
| D | 849 hCoV-19/South Africa/KRISP-0134/2020\|EPI_ISL_46748 | ···················· | ···················· | ···················· | ···················· | ···················· | ···················· |
| D | 850 hCoV-19/South Africa/KRISP-0136/2020\|EPI_ISL_46748 | ···················· | ···················· | ···················· | ···················· | ···················· | ···················· |
| D | 851 hCoV-19/South Africa/KRISP-0137/2020\|EPI_ISL_46748 | ···················· | ···················· | ···················· | ···················· | ···················· | ···················· |
| D | 856 hCoV-19/England/LIVE-9F6C8/2020\|EPI_ISL_472001\|202 | ···················· | ···················· | ···················· | ···················· | ···················· | ···················· |
| D | 857 hCoV-19/England/LIVE-9F6E6/2020\|EPI_ISL_472003\|202 | ···················· | ···················· | ···················· | ···················· | ···················· | ···················· |
| D | 858 hCoV-19/England/LIVE-9F6F5/2020\|EPI_ISL_472004\|202 | ···················· | ···················· | ···················· | ···················· | ···················· | ···················· |
| D | 859 hCoV-19/England/LIVE-9F701/2020\|EPI_ISL_472005\|202 | ···················· | ···················· | ···················· | ···················· | ···················· | ···················· |
| D | 860 hCoV-19/England/LIVE-9F710/2020\|EPI_ISL_472006\|202 | ···················· | ···················· | ···················· | ···················· | ···················· | ···················· |
| D | 861 hCoV-19/England/LIVE-9F72F/2020\|EPI_ISL_472007\|202 | ···················· | ···················· | ···················· | ···················· | ···················· | ···················· |
| D | 862 hCoV-19/England/LIVE-9F73E/2020\|EPI_ISL_472008\|202 | ···················· | ···················· | ···················· | ···················· | ···················· | ···················· |
| D | 863 hCoV-19/England/LIVE-9F74D/2020\|EPI_ISL_472009\|202 | ···················· | ···················· | ···················· | ···················· | ···················· | ···················· |
| D | 864 hCoV-19/England/LIVE-9F75C/2020\|EPI_ISL_472010\|202 | ···················· | ···················· | ···················· | ···················· | ···················· | ···················· |
| D | 865 hCoV-19/England/LIVE-9F76B/2020\|EPI_ISL_472011\|202 | ···················· | ···················· | ···················· | ···················· | ···················· | ···················· |
| D | 866 hCoV-19/England/LIVE-9F77A/2020\|EPI_ISL_472012\|202 | ···················· | ···················· | ···················· | ···················· | ···················· | ···················· |
| D | 867 hCoV-19/England/LIVE-9F789/2020\|EPI_ISL_472013\|202 | ···················· | ···················· | ···················· | ···················· | ···················· | ···················· |
| D | 868 hCoV-19/England/LIVE-9F798/2020\|EPI_ISL_472014\|202 | ···················· | ···················· | ···················· | ···················· | ···················· | ···················· |
| D | 869 hCoV-19/England/LIVE-9F7A7/2020\|EPI_ISL_472015\|202 | ···················· | ···················· | ···················· | ···················· | ···················· | ···················· |
| D | 870 hCoV-19/England/LIVE-9F7B6/2020\|EPI_ISL_472016\|202 | ···················· | ···················· | ···················· | ···················· | ···················· | ···················· |
| D | 871 hCoV-19/England/LIVE-9F84A/2020\|EPI_ISL_472018\|202 | ···················· | ···················· | ···················· | ···················· | ···················· | ···················· |
| D | 872 hCoV-19/England/LIVE-9F886/2020\|EPI_ISL_472019\|202 | ···················· | ···················· | ···················· | ···················· | ···················· | ···················· |
| D | 873 hCoV-19/England/LIVE-9F895/2020\|EPI_ISL_472020\|202 | ···················· | ···················· | ···················· | ···················· | ···················· | ···················· |
| D | 874 hCoV-19/England/LIVE-9F8A4/2020\|EPI_ISL_472021\|202 | ···················· | ···················· | ···················· | ···················· | ···················· | ···················· |
| D | 875 hCoV-19/England/LIVE-9F8C2/2020\|EPI_ISL_472022\|202 | ···················· | ···················· | ···················· | ···················· | ···················· | ···················· |
| D | 876 hCoV-19/Wales/PHWC-163486/2020\|EPI_ISL_473300\|2020 | ···················· | ···················· | ···················· | ···················· | ···················· | ···················· |
| D | 877 hCoV-19/Wales/PHWC-1634B3/2020\|EPI_ISL_473303\|2020 | ···················· | ···A················ | ···················· | ···················· | ···················· | ···················· |
| D | 878 hCoV-19/Wales/PHWC-1634E0/2020\|EPI_ISL_473305\|2020 | ···················· | ···················· | ···················· | ···················· | ···················· | ···················· |
| D | 879 hCoV-19/England/BIRM-5F08A/2020\|EPI_ISL_473306\|202 | ···················· | ···················· | ···················· | ···················· | ···················· | ···················· |
| D | 880 hCoV-19/England/BIRM-5F0A8/2020\|EPI_ISL_473308\|202 | ···················· | ···················· | ···················· | ···················· | ···················· | ···················· |
| D | 881 hCoV-19/England/BIRM-5F0D5/2020\|EPI_ISL_473310\|202 | ···················· | ···················· | ···················· | ···················· | ···················· | ···················· |
| D | 882 hCoV-19/England/BIRM-5F0E4/2020\|EPI_ISL_473311\|202 | ···················· | ···················· | ···················· | ···················· | ···················· | ···················· |
| D | 883 hCoV-19/England/BIRM-5F0F3/2020\|EPI_ISL_473312\|202 | ···················· | ···················· | ···················· | ···················· | ···················· | ···················· |
| D | 884 hCoV-19/England/BIRM-5F10F/2020\|EPI_ISL_473313\|202 | ···················· | ···················· | ···················· | ···················· | ···················· | ···················· |
| D | 885 hCoV-19/England/BIRM-5F11E/2020\|EPI_ISL_473314\|202 | ···················· | ···················· | ···················· | ···················· | ···················· | ···················· |
| D | 886 hCoV-19/England/BIRM-5F14B/2020\|EPI_ISL_473315\|202 | ···················· | ···················· | ···················· | ···················· | ···················· | ···················· |
| D | 887 hCoV-19/England/BIRM-5F15A/2020\|EPI_ISL_473316\|202 | ···················· | ···················· | ···················· | ···················· | ···················· | ···················· |
| D | 888 hCoV-19/England/BIRM-5F169/2020\|EPI_ISL_473317\|202 | ···················· | ···················· | ···················· | ···················· | ···················· | ···················· |
| D | 889 hCoV-19/England/BIRM-5F187/2020\|EPI_ISL_473318\|202 | ···················· | ···················· | ···················· | ···················· | ···················· | ···················· |
| D | 890 hCoV-19/England/BIRM-5F6B5/2020\|EPI_ISL_473319\|202 | ···················· | ···················· | ···················· | ···················· | ···················· | ···················· |
| D | 891 hCoV-19/England/BIRM-5F7C1/2020\|EPI_ISL_473332\|202 | ···················· | ···················· | ···················· | ···················· | ···················· | ···················· |
| D | 892 hCoV-19/England/BIRM-5F7D0/2020\|EPI_ISL_473333\|202 | ···················· | ···················· | ···················· | ···················· | ···················· | ···················· |
| D | 893 hCoV-19/England/BIRM-5F80A/2020\|EPI_ISL_473336\|202 | ···················· | ···················· | ···················· | ···················· | ···················· | ···················· |
| D | 894 hCoV-19/England/BIRM-5F837/2020\|EPI_ISL_473338\|202 | ···················· | ···················· | ···················· | ···················· | ···················· | ···················· |
| D | 895 hCoV-19/England/BIRM-5F855/2020\|EPI_ISL_473340\|202 | ···················· | ···················· | ···················· | ···················· | ···················· | ···················· |
| D | 896 hCoV-19/England/BIRM-5F882/2020\|EPI_ISL_473343\|202 | ···················· | ···················· | ···················· | ···················· | ···················· | ···················· |
| D | 897 hCoV-19/England/BIRM-5F891/2020\|EPI_ISL_473344\|202 | ···················· | ···················· | ···················· | ···················· | ···················· | ···················· |
| D | 899 hCoV-19/England/BIRM-5F8EC/2020\|EPI_ISL_473348\|202 | ···················· | ···················· | ···················· | ···················· | ···················· | ···················· |
| D | 900 hCoV-19/England/BIRM-5F8FB/2020\|EPI_ISL_473349\|202 | ···················· | ···················· | ···················· | ···················· | ···················· | ···················· |
| D | 901 hCoV-19/England/BIRM-5F916/2020\|EPI_ISL_473350\|202 | ···················· | ···················· | ···················· | ···················· | ···················· | ···················· |
| D | 902 hCoV-19/England/BIRM-5F934/2020\|EPI_ISL_473352\|202 | ···················· | ···················· | ···················· | ···················· | ···················· | ···················· |
| D | 903 hCoV-19/England/BIRM-5F970/2020\|EPI_ISL_473354\|202 | ···················· | ···················· | ···················· | ···················· | ···················· | ···················· |
| D | 904 hCoV-19/England/BIRM-5F99E/2020\|EPI_ISL_473356\|202 | ···················· | ···················· | ···················· | ···················· | ···················· | ···················· |
| D | 905 hCoV-19/England/BIRM-5F9AD/2020\|EPI_ISL_473357\|202 | ···················· | ···················· | ···················· | ···················· | ···················· | ···················· |
| D | 906 hCoV-19/England/BIRM-5F9BC/2020\|EPI_ISL_473358\|202 | ···················· | ···················· | ···················· | ···················· | ···················· | ···················· |
| D | 907 hCoV-19/England/BIRM-5F9DA/2020\|EPI_ISL_473360\|202 | ···················· | ···················· | ···················· | ···················· | ···················· | ···················· |
| D | 908 hCoV-19/England/BIRM-5F9E9/2020\|EPI_ISL_473361\|202 | ···················· | ···················· | ···················· | ···················· | ···················· | ···················· |
| D | 909 hCoV-19/England/BIRM-5F9F8/2020\|EPI_ISL_473362\|202 | ···················· | ···················· | ···················· | ···················· | ···················· | ···················· |
| D | 910 hCoV-19/England/BIRM-5FA22/2020\|EPI_ISL_473363\|202 | ···················· | ···················· | ···················· | ···················· | ···················· | ···················· |
| D | 911 hCoV-19/Wales/PHWC-35FAB/2020\|EPI_ISL_474600\|2020- | ···················· | ···················· | ···················· | ···················· | ···················· | ···················· |

| | | | | | | | | |
|---|---|---|---|---|---|---|---|---|
| D | 912 | hCoV-19/Wales/PHWC-36010/2020\|EPI_ISL_474607\|2020- | ···················· | ···················· | ···················· | ···················· | ···················· | ···················· |
| D | 913 | hCoV-19/Wales/PHWC-3602F/2020\|EPI_ISL_474608\|2020- | ···················· | ···················· | ···················· | ···················· | ···················· | ···················· |
| D | 914 | hCoV-19/Wales/PHWC-3603E/2020\|EPI_ISL_474609\|2020- | ···················· | ···················· | ···················· | ···················· | ···················· | ···················· |
| D | 915 | hCoV-19/Wales/PHWC-36098/2020\|EPI_ISL_474615\|2020- | ···················· | ····A··············· | ···················· | ···················· | ···················· | ···················· |
| D | 916 | hCoV-19/Wales/PHWC-360A7/2020\|EPI_ISL_474616\|2020- | ···················· | ···················· | ···················· | ···················· | ···················· | ···················· |
| D | 917 | hCoV-19/Wales/PHWC-3610E/2020\|EPI_ISL_474622\|2020- | ···················· | ···················· | ···················· | ···················· | ···················· | ···················· |
| D | 918 | hCoV-19/Wales/PHWC-3612C/2020\|EPI_ISL_474624\|2020- | ···················· | ···················· | ···················· | ···················· | ···················· | ···················· |
| D | 919 | hCoV-19/Wales/PHWC-3614A/2020\|EPI_ISL_474626\|2020- | ···················· | ···················· | ···················· | ···················· | ···················· | ···················· |
| D | 920 | hCoV-19/Wales/PHWC-361B3/2020\|EPI_ISL_474631\|2020- | ···················· | ···················· | ···················· | ···················· | ···················· | ···················· |
| D | 921 | hCoV-19/Wales/PHWC-361E0/2020\|EPI_ISL_474634\|2020- | ···················· | ···················· | ···················· | ···················· | ···················· | ···················· |
| D | 922 | hCoV-19/Wales/PHWC-3621A/2020\|EPI_ISL_474637\|2020- | ···················· | ···················· | ···················· | ···················· | ···················· | ···················· |
| D | 923 | hCoV-19/Wales/PHWC-36238/2020\|EPI_ISL_474639\|2020- | ···················· | ···················· | ···················· | ···················· | ···················· | ···················· |
| D | 924 | hCoV-19/Wales/PHWC-36308/2020\|EPI_ISL_474650\|2020- | ···················· | ···················· | ···················· | ···················· | ···················· | ···················· |
| D | 925 | hCoV-19/Wales/PHWC-36317/2020\|EPI_ISL_474651\|2020- | ···················· | ···················· | ···················· | ···················· | ···················· | ···················· |
| D | 926 | hCoV-19/Wales/PHWC-36405/2020\|EPI_ISL_474664\|2020- | ···················· | ···················· | ···················· | ···················· | ···················· | ···················· |
| D | 927 | hCoV-19/Wales/PHWC-3649C/2020\|EPI_ISL_474671\|2020- | ···················· | ···················· | ···················· | ···················· | ···················· | ···················· |
| D | 928 | hCoV-19/Wales/PHWC-364E7/2020\|EPI_ISL_474675\|2020- | ···················· | ···················· | ···················· | ···················· | ···················· | ···················· |
| D | 929 | hCoV-19/Wales/PHWC-364F6/2020\|EPI_ISL_474676\|2020- | ···················· | ···················· | ···················· | ···················· | ···················· | ···················· |
| D | 930 | hCoV-19/Wales/PHWC-36502/2020\|EPI_ISL_474677\|2020- | ···················· | ···················· | ···················· | ···················· | ···················· | ···················· |
| D | 931 | hCoV-19/Wales/PHWC-3654E/2020\|EPI_ISL_474680\|2020- | ···················· | ···················· | ···················· | ···················· | ···················· | ···················· |
| D | 932 | hCoV-19/Wales/PHWC-3656C/2020\|EPI_ISL_474682\|2020- | ···················· | ···················· | ···················· | ···················· | ···················· | ···················· |
| D | 933 | hCoV-19/Wales/PHWC-3658A/2020\|EPI_ISL_474684\|2020- | ···················· | ···················· | ···················· | ···················· | ···················· | ···················· |
| D | 934 | hCoV-19/Wales/PHWC-365B7/2020\|EPI_ISL_474686\|2020- | ···················· | ···················· | ···················· | ···················· | ···················· | ···················· |
| D | 935 | hCoV-19/Wales/PHWC-365F3/2020\|EPI_ISL_474689\|2020- | ···················· | ···················· | ···················· | ···················· | ···················· | ···················· |
| D | 936 | hCoV-19/Wales/PHWC-3662D/2020\|EPI_ISL_474692\|2020- | ···················· | ···················· | ···················· | ···················· | ···················· | ···················· |
| D | 937 | hCoV-19/Wales/PHWC-3663C/2020\|EPI_ISL_474693\|2020- | ···················· | ···················· | ···················· | ···················· | ···················· | ···················· |
| D | 938 | hCoV-19/Austria/CeMM0538/2020\|EPI_ISL_475900\|2020- | ···················· | ···················· | ···················· | ···················· | ···················· | ···················· |
| D | 939 | hCoV-19/Austria/CeMM0539/2020\|EPI_ISL_475901\|2020- | ···················· | ···················· | ···················· | ···················· | ···················· | ···················· |
| D | 940 | hCoV-19/Austria/CeMM0540/2020\|EPI_ISL_475902\|2020- | ···················· | ···················· | ···················· | ···················· | ···················· | ···················· |
| D | 941 | hCoV-19/Austria/CeMM0546/2020\|EPI_ISL_475903\|2020- | ···················· | ···················· | ···················· | ···················· | ···················· | ···················· |
| D | 942 | hCoV-19/Austria/CeMM0553/2020\|EPI_ISL_475904\|2020- | ···················· | ···················· | ···················· | ···················· | ···················· | ···················· |
| D | 943 | hCoV-19/Austria/CeMM0554/2020\|EPI_ISL_475905\|2020- | ···················· | ···················· | ···················· | ···················· | ···················· | ···················· |
| D | 944 | hCoV-19/Austria/CeMM0555/2020\|EPI_ISL_475906\|2020- | ···················· | ···················· | ···················· | ···················· | ···················· | ···················· |
| D | 945 | hCoV-19/Austria/CeMM0556/2020\|EPI_ISL_475907\|2020- | ···················· | ·············M······ | ···················· | ···················· | ···················· | ···················· |
| D | 946 | hCoV-19/Austria/CeMM0557/2020\|EPI_ISL_475908\|2020- | ···················· | ···················· | ···················· | ···················· | ···················· | ···················· |
| D | 947 | hCoV-19/Austria/CeMM0558/2020\|EPI_ISL_475909\|2020- | ···················· | ···················· | ···················· | ···················· | ···················· | ···················· |
| D | 948 | hCoV-19/Austria/CeMM0559/2020\|EPI_ISL_475910\|2020- | ···················· | ···················· | ···················· | ···················· | ···················· | ···················· |
| D | 949 | hCoV-19/Austria/CeMM0560/2020\|EPI_ISL_475911\|2020- | ···················· | ···················· | ···················· | ···················· | ···················· | ···················· |
| D | 950 | hCoV-19/Austria/CeMM0562/2020\|EPI_ISL_475912\|2020- | ···················· | ···················· | ···················· | ···················· | ···················· | ···················· |
| D | 951 | hCoV-19/Austria/CeMM0563/2020\|EPI_ISL_475913\|2020- | ···················· | ···················· | ···················· | ···················· | ···················· | ···················· |
| D | 952 | hCoV-19/Austria/CeMM0564/2020\|EPI_ISL_475914\|2020- | ···················· | ···················· | ···················· | ···················· | ···················· | ···················· |
| D | 953 | hCoV-19/Austria/CeMM0565/2020\|EPI_ISL_475915\|2020- | ···················· | ···················· | ···················· | ···················· | ···················· | R··················· |
| D | 954 | hCoV-19/Austria/CeMM0570/2020\|EPI_ISL_475919\|2020- | ···················· | ···················· | ···················· | ···················· | ···················· | ···················· |
| D | 955 | hCoV-19/Austria/CeMM0576/2020\|EPI_ISL_475923\|2020- | ···················· | ···················· | ···················· | ···················· | ···················· | ···················· |
| D | 956 | hCoV-19/Austria/CeMM0577/2020\|EPI_ISL_475924\|2020- | ···················· | ···················· | ···················· | ···················· | ···················· | ···················· |
| D | 957 | hCoV-19/Austria/CeMM0579/2020\|EPI_ISL_475926\|2020- | ···················· | ···················· | ···················· | ···················· | ···················· | ···················· |
| D | 958 | hCoV-19/Austria/CeMM0580/2020\|EPI_ISL_475927\|2020- | ···················· | ···················· | ···················· | ···················· | ···················· | ···················· |
| D | 959 | hCoV-19/Austria/CeMM0581/2020\|EPI_ISL_475928\|2020- | ···················· | ···················· | ···················· | ···················· | ···················· | ···················· |
| D | 960 | hCoV-19/Austria/CeMM0620/2020\|EPI_ISL_475929\|2020- | ···················· | ···················· | ···················· | ···················· | ···················· | ···················· |
| D | 961 | hCoV-19/Austria/CeMM0621/2020\|EPI_ISL_475930\|2020- | ···················· | ···················· | ···················· | ···················· | ···················· | ···················· |
| D | 962 | hCoV-19/Austria/CeMM0622/2020\|EPI_ISL_475931\|2020- | ···················· | ···················· | ···················· | ···················· | ···················· | ···················· |
| D | 963 | hCoV-19/Austria/CeMM0628/2020\|EPI_ISL_475932\|2020- | ···················· | ···················· | ···················· | ···················· | ···················· | ···················· |
| D | 964 | hCoV-19/Austria/CeMM0635/2020\|EPI_ISL_475934\|2020- | ···················· | ···················· | ···················· | ···················· | ···················· | ···················· |
| D | 965 | hCoV-19/Austria/CeMM0637/2020\|EPI_ISL_475935\|2020- | ···················· | ···················· | ···················· | ···················· | ···················· | ···················· |
| D | 966 | hCoV-19/Austria/CeMM0639/2020\|EPI_ISL_475936\|2020- | ···················· | ···················· | ···················· | ···················· | ···················· | ···················· |
| D | 967 | hCoV-19/Singapore/320/2020\|EPI_ISL_475937\|2020-05- | ···················· | ···················· | ···················· | ···················· | ···················· | ···················· |
| D | 968 | hCoV-19/Singapore/407/2020\|EPI_ISL_475938\|2020-04- | ···················· | ···················· | ···················· | ···················· | ···················· | ···················· |
| D | 969 | hCoV-19/Singapore/408/2020\|EPI_ISL_475939\|2020-04- | ···················· | ···················· | ···················· | ···················· | ···················· | ···················· |
| D | 970 | hCoV-19/Singapore/409/2020\|EPI_ISL_475940\|2020-04- | ···················· | ···················· | ···················· | ···················· | ···················· | ···················· |
| D | 971 | hCoV-19/Singapore/410/2020\|EPI_ISL_475941\|2020-04- | ···················· | ···················· | ···················· | ···················· | ···················· | ···················· |
| D | 972 | hCoV-19/Singapore/411/2020\|EPI_ISL_475942\|2020-04- | ···················· | ···················· | ···················· | ···················· | ···················· | ···················· |
| D | 973 | hCoV-19/Singapore/412/2020\|EPI_ISL_475943\|2020-05- | ···················· | ···················· | ···················· | ···················· | ···················· | ···················· |
| D | 974 | hCoV-19/Singapore/413/2020\|EPI_ISL_475944\|2020-04- | ···················· | ···················· | ···················· | ···················· | ···················· | ···················· |
| D | 975 | hCoV-19/Singapore/414/2020\|EPI_ISL_475945\|2020-04- | ···················· | ···················· | ···················· | ···················· | ···················· | ···················· |
| D | 976 | hCoV-19/Singapore/415/2020\|EPI_ISL_475946\|2020-04- | ···················· | ···················· | ···················· | ···················· | ···················· | ···R················ |
| D | 977 | hCoV-19/Singapore/416/2020\|EPI_ISL_475947\|2020-04- | ···················· | ···················· | ···················· | ···················· | ···················· | ················Y··· |
| D | 978 | hCoV-19/Singapore/417/2020\|EPI_ISL_475948\|2020-03- | ···················· | ···················· | ···················· | ···················· | ···················· | ···················· |
| D | 979 | hCoV-19/Singapore/418/2020\|EPI_ISL_475949\|2020-03- | ···················· | ···················· | ···················· | ···················· | ···················· | ···················· |
| D | 980 | hCoV-19/Singapore/419/2020\|EPI_ISL_475950\|2020-03- | ···················· | ···················· | ···················· | ···················· | ···················· | ···················· |
| D | 981 | hCoV-19/Singapore/420/2020\|EPI_ISL_475951\|2020-04- | ···················· | ···················· | ···················· | ···················· | ···················· | ···················· |
| D | 982 | hCoV-19/Singapore/422/2020\|EPI_ISL_475953\|2020-04- | ···················· | ···················· | ···················· | ···················· | ···················· | ···················· |
| D | 983 | hCoV-19/Singapore/423/2020\|EPI_ISL_475954\|2020-03- | ···················· | ···················· | ···················· | ···················· | ···················· | ···················· |
| D | 984 | hCoV-19/Singapore/424/2020\|EPI_ISL_475955\|2020-03- | ···················· | ···················· | ···················· | ···················· | ···················· | ···················· |
| D | 985 | hCoV-19/Singapore/426/2020\|EPI_ISL_475957\|2020-03- | ···················· | ···················· | ···················· | ···················· | ···················· | ···················· |
| D | 986 | hCoV-19/Singapore/427/2020\|EPI_ISL_475958\|2020-03- | ···················· | ···················· | ···················· | ···················· | ···················· | ···················· |
| D | 987 | hCoV-19/Singapore/428/2020\|EPI_ISL_475959\|2020-03- | ···················· | ···················· | ···················· | ···················· | ···················· | ···················· |
| D | 988 | hCoV-19/Singapore/429/2020\|EPI_ISL_475960\|2020-03- | ···················· | ···················· | ···················· | ···················· | ···················· | ···················· |
| D | 989 | hCoV-19/Singapore/431/2020\|EPI_ISL_475962\|2020-03- | ···················· | ···················· | ···················· | ···················· | ···················· | N··················· |

| | | | | | | | | | |
|---|---|---|---|---|---|---|---|---|---|
| D | 990 | hCoV-19/Singapore/432/2020\|EPI_ISL_475963\|2020-03- | ···················· | ···················· | ···················· | ···················· | ···················· | ···················· | |
| D | 991 | hCoV-19/Singapore/435/2020\|EPI_ISL_475966\|2020-03- | ···················· | ···················· | ···················· | ···················· | ···················· | ···················· | |
| D | 992 | hCoV-19/Singapore/436/2020\|EPI_ISL_475967\|2020-04- | ···················· | ···················· | ···················· | ···················· | ···················· | ···················· | |
| D | 993 | hCoV-19/Singapore/437/2020\|EPI_ISL_475968\|2020-03- | ···················· | ···················· | ···················· | ···················· | ···················· | ···················· | |
| D | 994 | hCoV-19/Singapore/438/2020\|EPI_ISL_475969\|2020-03- | ···················· | ···················· | ···················· | ···················· | ···················· | ···················· | |
| D | 995 | hCoV-19/Singapore/439/2020\|EPI_ISL_475970\|2020-03- | ···················· | ···················· | ···················· | ···················· | ···················· | ···················· | |
| D | 996 | hCoV-19/Singapore/440/2020\|EPI_ISL_475971\|2020-04- | ···················· | ···················· | ···················· | ···················· | ···················· | ···················· | |
| D | 997 | hCoV-19/Singapore/441/2020\|EPI_ISL_475972\|2020-04- | ···················· | ···················· | ···················· | ···················· | ···················· | ···················· | |
| D | 998 | hCoV-19/Singapore/442/2020\|EPI_ISL_475973\|2020-04- | ···················· | ···················· | ···················· | ···················· | ···················· | ···················· | |
| D | 999 | hCoV-19/Singapore/448/2020\|EPI_ISL_475979\|2020-04- | ···················· | ···················· | ···················· | ···················· | ···················· | ···················· | |
| D | 1000 | hCoV-19/Singapore/450/2020\|EPI_ISL_475981\|2020-03- | ···················· | ···················· | ···················· | ···················· | ···················· | ···················· | |
| D | 1001 | hCoV-19/Singapore/464/2020\|EPI_ISL_475995\|2020-05- | ···················· | ···················· | ···················· | ···················· | ···················· | ···················· | |
| D | 1002 | hCoV-19/Singapore/466/2020\|EPI_ISL_475997\|2020-01- | ···················· | ···················· | ···················· | ···················· | ···················· | ···················· | |
| D | 1003 | hCoV-19/Scotland/CVR3813/2020\|EPI_ISL_478139\|2020- | ···················· | ···················· | ···················· | ···················· | ···················· | ···················· | |
| D | 1004 | hCoV-19/Scotland/CVR3814/2020\|EPI_ISL_478140\|2020- | ···················· | ···················A | ···················· | ···················· | ···················· | ···················· | 1 |
| D | 1005 | hCoV-19/Scotland/CVR3816/2020\|EPI_ISL_478142\|2020- | ···················· | ···················· | ···················· | ···················· | ···················· | ···················· | |
| D | 1006 | hCoV-19/Scotland/CVR3817/2020\|EPI_ISL_478143\|2020- | ···················· | ···················· | ···················· | ···················· | ···················· | ···················· | |
| D | 1007 | hCoV-19/Scotland/CVR3818/2020\|EPI_ISL_478144\|2020- | ···················· | ···················A | ···················· | ···················· | ···················· | ···················· | 1 |
| D | 1008 | hCoV-19/Scotland/CVR3820/2020\|EPI_ISL_478146\|2020- | ···················· | ···················A | ···················· | ···················· | ···················· | ···················· | 1 |
| D | 1009 | hCoV-19/Scotland/CVR3821/2020\|EPI_ISL_478147\|2020- | ···················· | ···················A | ···················· | ···················· | ···················· | ···················· | 1 |
| D | 1010 | hCoV-19/Scotland/CVR3822/2020\|EPI_ISL_478148\|2020- | ···················· | ···················· | ···················· | ···················· | ···················· | ···················· | |
| D | 1011 | hCoV-19/Scotland/CVR3823/2020\|EPI_ISL_478149\|2020- | ···················· | ···················A | ···················· | ···················· | ···················· | ···················· | 1 |
| D | 1012 | hCoV-19/Scotland/EDB4672/2020\|EPI_ISL_478180\|2020- | ···················· | ···················· | ···················· | ···················· | ···················· | ···················· | |
| D | 1013 | hCoV-19/Scotland/EDB4680/2020\|EPI_ISL_478182\|2020- | ···················· | ·A·················· | ···················· | ···················· | ···················· | ···················· | |
| D | 1014 | hCoV-19/Scotland/EDB4681/2020\|EPI_ISL_478183\|2020- | ···················· | ·A·················· | ···················· | ···················· | ···················· | ···················· | |
| D | 1015 | hCoV-19/Scotland/EDB4690/2020\|EPI_ISL_478184\|2020- | ···················· | ···················· | ···················· | ···················· | ···················· | ···················· | |
| D | 1016 | hCoV-19/Scotland/EDB4777/2020\|EPI_ISL_478186\|2020- | ···················· | ·A·················· | ···················· | ···················· | ···················· | ···················· | |
| D | 1017 | hCoV-19/Scotland/EDB4807/2020\|EPI_ISL_478188\|2020- | ···················· | ·A·················· | ···················· | ···················· | ···················· | ···················· | |
| D | 1018 | hCoV-19/Scotland/EDB4859/2020\|EPI_ISL_478190\|2020- | ···················· | ···················· | ···················· | ···················· | ···················· | ···················· | |
| D | 1019 | hCoV-19/Scotland/EDB4869/2020\|EPI_ISL_478192\|2020- | ···················· | ···················· | ···················· | ···················· | ···················· | ···················· | |
| D | 1020 | hCoV-19/Scotland/EDB4871/2020\|EPI_ISL_478193\|2020- | ···················· | ···················· | ···················· | ···················· | ···················· | ···················· | |
| D | 1021 | hCoV-19/Scotland/EDB4873/2020\|EPI_ISL_478194\|2020- | ···················· | ···················· | ···················· | ···················· | ···················· | ···················· | |
| D | 1022 | hCoV-19/Scotland/EDB4898/2020\|EPI_ISL_478195\|2020- | ···················· | ···················· | ···················· | ···················· | ···················· | ···················· | |
| D | 1023 | hCoV-19/Scotland/EDB6057/2020\|EPI_ISL_478199\|2020- | ···················· | ·A·················· | ···················· | ···················· | ···················· | ···················· | |
| D | 1024 | hCoV-19/Germany/MV-Tv4-20-Tier-4/2020\|EPI_ISL_4911 | ···················· | ···················· | ···················· | ···················· | ···················· | ···················· | |
| D | 1025 | hCoV-19/Singapore/280/2020\|EPI_ISL_462398\|2020-04- | ···················· | ···················· | ···················· | ···················· | ···················· | ···················· | |
| D | 1026 | hCoV-19/Singapore/281/2020\|EPI_ISL_462399\|2020-04- | ···················· | ···················· | ···················· | ···················· | ···················· | ···················· | |
| D | 1027 | hCoV-19/Shaoxing/09/2020\|EPI_ISL_463901\|2020-02-05 | ···················· | ···················· | ···················· | ···················· | ···········N········ | ···················· | |
| D | 1028 | hCoV-19/Canada/QC_AC6.1/2020\|EPI_ISL_463904\|2020-0 | ···················· | ···················· | ···················· | ···················· | ···················· | ···················· | |
| D | 1030 | hCoV-19/Canada/QC_AE6/2020\|EPI_ISL_463906\|2020-04- | ···················· | ···················· | ···················· | ···················· | ···················· | ···················· | |
| D | 1031 | hCoV-19/Canada/QC_AE9/2020\|EPI_ISL_463908\|2020-04- | ···················· | ···················· | ···················· | ···················· | ···················· | ···················· | |
| D | 1032 | hCoV-19/Canada/QC_AF1/2020\|EPI_ISL_463909\|2020-04- | ···················· | ···················· | ···················· | ···················· | ···················· | ···················· | |
| D | 1033 | hCoV-19/Canada/QC_AF2/2020\|EPI_ISL_463910\|2020-04- | ···················· | ···················· | ···················· | ···················· | ···················· | ···················· | |
| D | 1036 | hCoV-19/Canada/QC_AH2/2020\|EPI_ISL_463914\|2020-04- | ···················· | ···················· | ···················· | ···················· | ···················· | ···················· | |
| D | 1037 | hCoV-19/Canada/QC_AI5/2020\|EPI_ISL_463917\|2020-04- | ···················· | ···················· | ···················· | ···················· | ···················· | ···················· | |
| D | 1038 | hCoV-19/Canada/QC_AI9/2020\|EPI_ISL_463918\|2020-04- | ···················· | ···················· | ···················· | ···················· | ···················· | ···················· | |
| D | 1039 | hCoV-19/Canada/QC_AK5/2020\|EPI_ISL_463919\|2020-04- | ···················· | ···················· | ···················· | ···················· | ···················· | ···················· | |
| D | 1040 | hCoV-19/England/20149006704/2020\|EPI_ISL_466120\|20 | ···················· | ···················· | ···················· | ···················· | ···················· | ···················· | |
| D | 1041 | hCoV-19/England/20149006904/2020\|EPI_ISL_466122\|20 | ···················· | ···················· | ···················· | ···················· | ···················· | ···················· | |
| D | 1042 | hCoV-19/England/20149007104/2020\|EPI_ISL_466124\|20 | ···················· | ···················· | ···················· | ···················· | ···················· | ···················· | |
| D | 1043 | hCoV-19/England/20149007604/2020\|EPI_ISL_466129\|20 | ···················· | ···················· | ···················· | ···················· | ···················· | ···················· | |
| D | 1044 | hCoV-19/England/20149007904/2020\|EPI_ISL_466131\|20 | ···················· | ···················· | ···················· | ···················· | ···················· | ·····M·············· | |
| D | 1045 | hCoV-19/England/20149008004/2020\|EPI_ISL_466132\|20 | ···················· | ···················· | ···················· | ···················· | ···················· | ···················· | |
| D | 1048 | hCoV-19/England/20149008704/2020\|EPI_ISL_466139\|20 | ···················· | ·A·················· | ···················· | ···················· | ···················· | ···················· | |
| D | 1049 | hCoV-19/England/20149016104/2020\|EPI_ISL_466161\|20 | ···················· | ···················· | ···················· | ···················· | ···················· | ···················· | |
| D | 1050 | hCoV-19/England/20149021404/2020\|EPI_ISL_466162\|20 | ···················· | ···················· | ···················· | ···················· | ···················· | ···················· | |
| D | 1051 | hCoV-19/England/20149021704/2020\|EPI_ISL_466163\|20 | ···················· | ···················· | ···················· | ···················· | ···················· | ···················· | |
| D | 1053 | hCoV-19/England/20149022004/2020\|EPI_ISL_466165\|20 | ···················· | ···················· | ···················· | ···················· | ·NNN················ | ···················· | 1 |
| D | 1054 | hCoV-19/England/20149022204/2020\|EPI_ISL_466166\|20 | ···················· | ···················· | ···················· | ···················· | ···················· | ···················· | |
| D | 1055 | hCoV-19/England/20149022304/2020\|EPI_ISL_466167\|20 | ···················· | ···················· | ···················· | ···················· | ···················· | ···················· | |
| D | 1056 | hCoV-19/England/20149022404/2020\|EPI_ISL_466168\|20 | ···················· | ···················· | ···················· | ···················· | ···················· | ···················· | |
| D | 1057 | hCoV-19/England/20149022504/2020\|EPI_ISL_466169\|20 | ···················· | ···················· | ···················· | ···················· | ···················· | ···················· | |
| D | 1059 | hCoV-19/England/20149023904/2020\|EPI_ISL_466173\|20 | ···················· | ···················· | ···················· | ···················· | ···················· | ···················· | |
| D | 1061 | hCoV-19/England/20149025404/2020\|EPI_ISL_466176\|20 | ···················· | ···················· | ···················· | ···················· | ···················· | ···················· | |
| D | 1062 | hCoV-19/England/20149026204/2020\|EPI_ISL_466178\|20 | ···················· | ···················· | ···················· | ···················· | ···················· | ···················· | |
| D | 1063 | hCoV-19/England/20149027404/2020\|EPI_ISL_466180\|20 | ···················· | ···················· | ···················· | ···················· | ···················· | ···················· | |
| D | 1066 | hCoV-19/England/20149035304/2020\|EPI_ISL_466186\|20 | ···················· | ···················· | ···················· | ···················· | ···················· | ···················· | |
| D | 1068 | hCoV-19/England/20149041204/2020\|EPI_ISL_466191\|20 | ···················· | ···················· | ···················· | ···················· | ···················· | ···················· | |
| D | 1070 | hCoV-19/England/20149043704/2020\|EPI_ISL_466195\|20 | ···················· | ···················· | ···················· | ···················· | ···················· | ···················· | |
| D | 1071 | hCoV-19/England/20149043804/2020\|EPI_ISL_466196\|20 | ···················· | ···················· | ···················· | ···················· | ···················· | ···················· | |
| D | 1074 | hCoV-19/England/20149044504/2020\|EPI_ISL_466199\|20 | ···················· | ···················· | ···················· | ···················· | ···················· | ···················· | |
| D | 1075 | hCoV-19/Spain/MD-IBV-003684/2020\|EPI_ISL_467200\|20 | ···················· | ···················· | ···················· | ···················· | ···················· | ···················· | |
| D | 1078 | hCoV-19/South Africa/KRISP-0065/2020\|EPI_ISL_46744 | ···················· | ···················· | ···················· | ···················· | ···················· | ···················· | |
| D | 1079 | hCoV-19/South Africa/KRISP-0066/2020\|EPI_ISL_46745 | ···················· | ···················· | ···················· | ···················· | ···················· | ···················· | |
| D | 1080 | hCoV-19/South Africa/KRISP-0067/2020\|EPI_ISL_46745 | ···················· | ···················· | ···················· | ···················· | ···················· | ···················· | |
| D | 1081 | hCoV-19/South Africa/KRISP-0075/2020\|EPI_ISL_46745 | ···················· | ···················· | ···················· | ···················· | ···················· | ···················· | |
| D | 1082 | hCoV-19/South Africa/KRISP-0080/2020\|EPI_ISL_46745 | ···················· | ···················· | ···················· | ···················· | ···················· | ···················· | |
| D | 1083 | hCoV-19/South Africa/KRISP-0081/2020\|EPI_ISL_46745 | ···················· | ···················· | ···················· | ···················· | ···················· | ···················· | |

```
D 1084 hCoV-19/South Africa/KRISP-0084/2020|EPI_ISL_46745 ···················· ···················· ···················· ···················· ···················· ····················
D 1085 hCoV-19/South Africa/KRISP-0088/2020|EPI_ISL_46745 ···················· ···················· ···················· ···················· ···················· ····················
D 1086 hCoV-19/South Africa/KRISP-0091/2020|EPI_ISL_46746 ···················· ···················· ···················· ···················· ···················· ····················
D 1087 hCoV-19/South Africa/KRISP-0096/2020|EPI_ISL_46746 ···················· ···················· ···················· ···················· ···················· ····················
D 1088 hCoV-19/South Africa/KRISP-0097/2020|EPI_ISL_46746 ···················· ···················· ···················· ···················· ···················· ····················
D 1089 hCoV-19/South Africa/KRISP-0111/2020|EPI_ISL_46746 ···················· ···················· ···················· ···················· ···················· ····················
D 1090 hCoV-19/South Africa/KRISP-0112/2020|EPI_ISL_46746 ···················· ···················· ···················· ···················· ···················· ····················
D 1091 hCoV-19/South Africa/KRISP-0113/2020|EPI_ISL_46746 ···················· ···················· ···················· ···················· ···················· ····················
D 1092 hCoV-19/South Africa/KRISP-0115/2020|EPI_ISL_46746 ···················· ···················· ···················· ···················· ···················· ····················
D 1093 hCoV-19/South Africa/KRISP-0116/2020|EPI_ISL_46746 ···················· ···················· ···················· ···················· ···················· ····················
D 1094 hCoV-19/South Africa/KRISP-0138/2020|EPI_ISL_46749 ···················· ···················· ···················· ···················· ···················· ····················
D 1095 hCoV-19/South Africa/KRISP-0139/2020|EPI_ISL_46749 ···················· ···················· ···················· ···················· ···················· ····················
D 1096 hCoV-19/South Africa/KRISP-0143/2020|EPI_ISL_46749 ···················· ···················· ···················· ···················· ···················· ····················
D 1097 hCoV-19/South Africa/KRISP-0144/2020|EPI_ISL_46749 ···················· ···················· ···················· ···················· ···················· ····················
D 1098 hCoV-19/South Africa/KRISP-0145/2020|EPI_ISL_46749 ···················· ···················· ···················· ···················· ···················· ····················
D 1099 hCoV-19/South Africa/KRISP-0146/2020|EPI_ISL_46749 ···················· ···················· ···················· ···················· ···················· ····················
D 1100 hCoV-19/South Africa/KRISP-0147/2020|EPI_ISL_46749 ···················· ···················· ···················· ···················· ···················· ····················
D 1101 hCoV-19/South Africa/KRISP-0148/2020|EPI_ISL_46749 ···················· ···················· ···················· ···················· ···················· ····················
D 1102 hCoV-19/South Africa/KRISP-0149/2020|EPI_ISL_46749 ···················· ···················· ···················· ···················· ···················· ····················
D 1103 hCoV-19/USA/CA-CZB-1585/2020|EPI_ISL_468515|2020-0 ···················· ···················· ···················· ···················· ···················· ····················
D 1104 hCoV-19/USA/CA-CZB-1587/2020|EPI_ISL_468517|2020-0 ···················· ···················· ···················· ···················· ···················· ····················
D 1105 hCoV-19/USA/CA-CZB-1588/2020|EPI_ISL_468518|2020-0 ···················· ···················· ···················· ···················· ···················· ····················
D 1106 hCoV-19/USA/CA-CZB-1589/2020|EPI_ISL_468519|2020-0 ···················· ··············C····· ···················· ···················· ···················· ····················
D 1107 hCoV-19/USA/CA-CZB-1590/2020|EPI_ISL_468520|2020-0 ···················· ···················· ···················· ···················· ···················· ····················
D 1108 hCoV-19/USA/CA-CZB-1591/2020|EPI_ISL_468521|2020-0 ···················· ···················· ···················· ···················· ···················· ····················
D 1109 hCoV-19/USA/CA-CZB-1592/2020|EPI_ISL_468522|2020-0 ···················· ···················· ···················· ···················· ···················· ····················
D 1110 hCoV-19/USA/CA-CZB-1593/2020|EPI_ISL_468523|2020-0 ···················· ···················· ···················· ···················· ···················· ····················
D 1111 hCoV-19/USA/CA-CZB-1594/2020|EPI_ISL_468524|2020-0 ···················· ···················· ···················· ···················· ···················· ····················
D 1112 hCoV-19/USA/CA-CZB-1596/2020|EPI_ISL_468525|2020-0 ···················· ···················· ···················· ···················· ···················· ····················
D 1113 hCoV-19/USA/CA-CZB-1597/2020|EPI_ISL_468526|2020-0 ···················· ···················· ···················· ···················· ···················· ····················
D 1114 hCoV-19/USA/CA-CZB-1598/2020|EPI_ISL_468527|2020-0 ···················· ···················· ···················· ···················· ···················· ····················
D 1115 hCoV-19/USA/CA-CZB-1599/2020|EPI_ISL_468528|2020-0 ···················· ···················· ···················· ···················· ···················· ····················
D 1116 hCoV-19/USA/CA-CZB-1600/2020|EPI_ISL_468529|2020-0 ···················· ···················· ···················· ···················· ···················· ····················
D 1117 hCoV-19/USA/CA-CZB-1601/2020|EPI_ISL_468530|2020-0 ···················· ···················· ···················· ···················· ···················· ····················
D 1118 hCoV-19/USA/CA-CZB-1602/2020|EPI_ISL_468531|2020-0 ···················· ···················· ···················· ···················· ···················· ····················
D 1119 hCoV-19/USA/CA-CZB-1603/2020|EPI_ISL_468532|2020-0 ···················· ···················· ···················· ···················· ···················· ····················
D 1120 hCoV-19/USA/CA-CZB-1604/2020|EPI_ISL_468533|2020-0 ···················· ···················· ···················· ···················· ···················· ····················
D 1121 hCoV-19/USA/CA-CZB-1605/2020|EPI_ISL_468534|2020-0 ···················· ···················· ···················· ···················· ···················· ····················
D 1122 hCoV-19/USA/CA-CZB-1606/2020|EPI_ISL_468535|2020-0 ···················· ···················· ···················· ···················· ···················· ····················
D 1124 hCoV-19/Spain/AR-IBV-2472/2020|EPI_ISL_468779|2020 ···················· ···················· ···················· ···················· ·········A·········· ····················
D 1126 hCoV-19/Spain/AR-IBV-2461/2020|EPI_ISL_468781|2020 ···················· ···················· ···················· ···················· ···················· ····················
D 1142 hCoV-19/England/BRIS-128308/2020|EPI_ISL_469801|20 ···················· ···················· ···················· ···················· ···················· ····················
D 1143 hCoV-19/England/BRIS-127B2F/2020|EPI_ISL_469802|20 ···················· ···················· ···················· ···················· ···················· ····················
D 1144 hCoV-19/England/BRIS-127E17/2020|EPI_ISL_469803|20 ···················· ···················· ···················· ···················· ···················· ····················
D 1145 hCoV-19/England/BRIS-1270C6/2020|EPI_ISL_469804|20 ···················· ···················· ···················· ···················· ···················· ····················
D 1146 hCoV-19/England/BRIS-12B129/2020|EPI_ISL_469809|20 ···················· ···················· ···················· ···················· ···················· ····················
D 1147 hCoV-19/England/BRIS-12B29F/2020|EPI_ISL_469810|20 ···················· ···················· ···················· ···················· ···················· ····················
D 1148 hCoV-19/England/BRIS-12B156/2020|EPI_ISL_469811|20 ···················· ···················· ···················· ···················· ···················· ····················
D 1149 hCoV-19/England/BRIS-12B095/2020|EPI_ISL_469814|20 ···················· ···················· ···················· ···················· ···················· ····················
D 1150 hCoV-19/England/BRIS-12AFC6/2020|EPI_ISL_469818|20 ···················· ···················· ···················· ···················· ···················· ····················
D 1151 hCoV-19/England/BRIS-12B077/2020|EPI_ISL_469819|20 ···················· ···················· ···················· ···················· ···················· ····················
D 1152 hCoV-19/England/BRIS-12B086/2020|EPI_ISL_469820|20 ···················· ···················· ···················· ···················· ···················· ····················
D 1153 hCoV-19/England/BRIS-12B262/2020|EPI_ISL_469822|20 ···················· ···················· ···················· ···················· ···················· ····················
D 1154 hCoV-19/England/BRIS-12B147/2020|EPI_ISL_469840|20 ···················· ···················· ···················· ···················· ···················· ····················
D 1155 hCoV-19/Northern Ireland/NIRE-101754/2020|EPI_ISL_ ···················· ···················· ···················· ···················· ···················· ····················
D 1156 hCoV-19/Northern Ireland/NIRE-101736/2020|EPI_ISL_ ···················· ···················· ···················· ···················· ···················· ····················
D 1157 hCoV-19/Northern Ireland/NIRE-100EE7/2020|EPI_ISL_ ···················· ···················· ···················· ···················· ···················· ····················
D 1158 hCoV-19/Northern Ireland/NIRE-1024C5/2020|EPI_ISL_ ···················· ···················· ···················· ···················· ···················· ····················
D 1159 hCoV-19/Northern Ireland/NIRE-FF81B/2020|EPI_ISL_4 ···················· ···················· ···················· ···················· ···················· ····················
D 1160 hCoV-19/Northern Ireland/NIRE-FF2A4/2020|EPI_ISL_4 ···················· ···················· ···················· ···················· ···················· ····················
D 1161 hCoV-19/Northern Ireland/NIRE-FF65D/2020|EPI_ISL_4 ···················· ···················· ···················· ···················· ···················· ····················
D 1162 hCoV-19/Northern Ireland/NIRE-101280/2020|EPI_ISL_ ···················· ···················· ···················· ···················· ···················· ····················
D 1163 hCoV-19/Northern Ireland/NIRE-102708/2020|EPI_ISL_ ···················· ···················· ···················· ···················· ···················· ····················
D 1164 hCoV-19/Northern Ireland/NIRE-FF6D5/2020|EPI_ISL_4 ···················· ···················· ···················· ···················· ···················· ····················
D 1165 hCoV-19/Northern Ireland/NIRE-10261A/2020|EPI_ISL_ ···················· ···················· ···················· ···················· ···················· ····················
D 1166 hCoV-19/Northern Ireland/NIRE-FF523/2020|EPI_ISL_4 ···················· ···················· ···················· ···················· ···················· ····················
D 1167 hCoV-19/Northern Ireland/NIRE-101244/2020|EPI_ISL_ ···················· ···················· ···················· ···················· ···················· ····················
D 1168 hCoV-19/Northern Ireland/NIRE-102A3C/2020|EPI_ISL_ ···················· ···················· ···················· ···················· ···················· ····················
D 1169 hCoV-19/Northern Ireland/NIRE-1028F6/2020|EPI_ISL_ ···················· ···················· ···················· ···················· ···················· ····················
D 1170 hCoV-19/Northern Ireland/NIRE-102823/2020|EPI_ISL_ ···················· ···················· ···················· ···················· ···················· ····················
D 1171 hCoV-19/Northern Ireland/NIRE-100CB0/2020|EPI_ISL_ ···················· ···················· ···················· ···················· ···················· ····················
D 1172 hCoV-19/Australia/WA12/2020|EPI_ISL_470830|2020-03 ···················· ···················· ···················· ···················· ···················· ····················
D 1173 hCoV-19/Australia/WA13/2020|EPI_ISL_470831|2020-04 ···················· ···················· ···················· ···················· ···················· ····················
D 1174 hCoV-19/Australia/WA15/2020|EPI_ISL_470833|2020-04 ···················· ···················· ···················· ···················· ···················· ····················
D 1175 hCoV-19/Australia/WA17/2020|EPI_ISL_470835|2020-04 ···················· ···················· ···················· ···················· ···················· ····················
D 1176 hCoV-19/Australia/WA21/2020|EPI_ISL_470839|2020-04 ···················· ···················· ···················· ···················· ···················· ····················
D 1177 hCoV-19/Australia/WA22/2020|EPI_ISL_470840|2020-03 ···················· ···················· ···················· ···················· ···················· ····················
D 1178 hCoV-19/Australia/WA23/2020|EPI_ISL_470841|2020-03 ···················· ···················· ···················· ···················· ···················· ····················
```

| | | | | | | | | | |
|---|---|---|---|---|---|---|---|---|---|
| D | 1179 | hCoV-19/Australia/WA25/2020\|EPI_ISL_470843\|2020-04 | .................... | .................... | .................... | .................... | .................... | .................... | |
| D | 1180 | hCoV-19/Australia/WA28/2020\|EPI_ISL_470845\|2020-03 | .................... | .................... | .................... | .................... | .................... | .................... | |
| D | 1181 | hCoV-19/Australia/WA29/2020\|EPI_ISL_470846\|2020-03 | .................... | .................... | .................... | .................... | .................... | .................... | |
| D | 1182 | hCoV-19/Australia/WA30/2020\|EPI_ISL_470847\|2020-03 | .................... | .................... | .................... | .................... | .................... | .................... | |
| D | 1183 | hCoV-19/Australia/WA31/2020\|EPI_ISL_470848\|2020-03 | .................... | .................... | .................... | .................... | .................... | .................... | |
| D | 1184 | hCoV-19/Australia/WA32/2020\|EPI_ISL_470849\|2020-03 | .................... | .................... | .................... | .................... | .................... | .................... | |
| D | 1185 | hCoV-19/Wales/PHWC-160D60/2020\|EPI_ISL_473046\|2020 | .................... | .................... | .................... | .................... | .................... | .................... | |
| D | 1186 | hCoV-19/Wales/PHWC-160D8E/2020\|EPI_ISL_473048\|2020 | .................... | .................... | .................... | .................... | .................... | .................... | |
| D | 1187 | hCoV-19/Wales/PHWC-160DBB/2020\|EPI_ISL_473051\|2020 | .................... | .................... | .................... | .................... | .................... | .................... | |
| D | 1188 | hCoV-19/Wales/PHWC-160DCA/2020\|EPI_ISL_473052\|2020 | .................... | .................... | .................... | .................... | .................... | .................... | |
| D | 1189 | hCoV-19/Wales/PHWC-160DD9/2020\|EPI_ISL_473053\|2020 | .................... | .................... | .................... | .................... | .................... | .................... | |
| D | 1190 | hCoV-19/Wales/PHWC-160DF7/2020\|EPI_ISL_473055\|2020 | .................... | .................... | .................... | .................... | .................... | .................... | |
| D | 1191 | hCoV-19/Wales/PHWC-160E21/2020\|EPI_ISL_473056\|2020 | .................... | .................... | .................... | .................... | .................... | .................... | |
| D | 1192 | hCoV-19/Wales/PHWC-160E5E/2020\|EPI_ISL_473059\|2020 | .................... | .................... | .................... | .................... | .................... | .................... | |
| D | 1193 | hCoV-19/Wales/PHWC-162584/2020\|EPI_ISL_473090\|2020 | .................... | .................... | .................... | .................... | .................... | .................... | |
| D | 1194 | hCoV-19/Wales/PHWC-1625A2/2020\|EPI_ISL_473092\|2020 | .................... | ....A............... | .................... | .................... | .................... | .................... | |
| D | 1195 | hCoV-19/Wales/PHWC-1625DF/2020\|EPI_ISL_473095\|2020 | .................... | .................... | .................... | .................... | .................... | .................... | |
| D | 1196 | hCoV-19/Wales/PHWC-1625FD/2020\|EPI_ISL_473097\|2020 | .................... | .................... | .................... | .................... | .................... | .................... | |
| D | 1197 | hCoV-19/Wales/PHWC-329CE/2020\|EPI_ISL_474377\|2020- | .................... | .................... | .................... | .................... | .................... | .................... | |
| D | 1198 | hCoV-19/Wales/PHWC-329DD/2020\|EPI_ISL_474378\|2020- | .................... | .................... | .................... | .................... | .................... | .................... | |
| D | 1199 | hCoV-19/Wales/PHWC-32A16/2020\|EPI_ISL_474382\|2020- | .................... | .................... | .................... | .................... | .................... | .................... | |
| D | 1200 | hCoV-19/Wales/PHWC-32A34/2020\|EPI_ISL_474384\|2020- | .................... | .................... | .................... | .................... | .................... | .................... | |
| D | 1201 | hCoV-19/Wales/PHWC-32A61/2020\|EPI_ISL_474387\|2020- | .................... | .A.................. | .................... | .................... | .................... | .................... | |
| D | 1202 | hCoV-19/Wales/PHWC-32A70/2020\|EPI_ISL_474388\|2020- | .................... | .................... | .................... | .................... | .................... | .................... | |
| D | 1203 | hCoV-19/Wales/PHWC-32AAD/2020\|EPI_ISL_474391\|2020- | .................... | .................... | .................... | .................... | .................... | .................... | |
| D | 1204 | hCoV-19/Wales/PHWC-32AE9/2020\|EPI_ISL_474395\|2020- | .................... | .................... | .................... | .................... | .................... | .................... | |
| D | 1205 | hCoV-19/Wales/PHWC-32B04/2020\|EPI_ISL_474397\|2020- | .................... | .................... | .................... | .................... | .................... | .................... | |
| D | 1206 | hCoV-19/Wales/PHWC-32B13/2020\|EPI_ISL_474398\|2020- | .................... | ....A............... | .................... | .................... | .................... | .................... | |
| D | 1208 | hCoV-19/Scotland/CVR3773/2020\|EPI_ISL_478107\|2020- | .................... | .................... | .................... | .................... | .................... | .................... | |
| D | 1209 | hCoV-19/Scotland/CVR3774/2020\|EPI_ISL_478108\|2020- | .................... | .................... | .................... | .................... | .................... | .................... | |
| D | 1210 | hCoV-19/Scotland/CVR3776/2020\|EPI_ISL_478109\|2020- | .................... | .................... | .................... | .................... | .................... | .................... | |
| D | 1211 | hCoV-19/Scotland/CVR3779/2020\|EPI_ISL_478112\|2020- | .................... | .................... | .................... | .................... | .................... | .................... | |
| D | 1212 | hCoV-19/Scotland/CVR3781/2020\|EPI_ISL_478114\|2020- | .................... | .................... | .................... | .................... | .................... | .................... | |
| D | 1213 | hCoV-19/Scotland/CVR3782/2020\|EPI_ISL_478115\|2020- | .................... | .................... | .................... | .................... | .................... | .................... | |
| D | 1214 | hCoV-19/Scotland/CVR3788/2020\|EPI_ISL_478116\|2020- | .................... | ...................A | .................... | .................... | .................... | .................... | 1 |
| D | 1215 | hCoV-19/Scotland/CVR3789/2020\|EPI_ISL_478117\|2020- | .................... | .................... | .................... | .................... | .................... | .................... | |
| D | 1216 | hCoV-19/Scotland/CVR3790/2020\|EPI_ISL_478118\|2020- | .................... | ...................A | .................... | .................... | .................... | .................... | 1 |
| D | 1217 | hCoV-19/Scotland/CVR3791/2020\|EPI_ISL_478119\|2020- | .................... | .................... | .................... | .................... | .................... | .................... | |
| D | 1218 | hCoV-19/Scotland/CVR3793/2020\|EPI_ISL_478121\|2020- | .................... | .................... | .................... | .................... | .................... | .................... | |
| D | 1219 | hCoV-19/Scotland/CVR3794/2020\|EPI_ISL_478122\|2020- | .................... | ...................A | .................... | .................... | .................... | .................... | 1 |
| D | 1220 | hCoV-19/Scotland/CVR3796/2020\|EPI_ISL_478124\|2020- | .................... | .................... | .................... | .................... | .................... | .................... | |
| D | 1221 | hCoV-19/Scotland/CVR3797/2020\|EPI_ISL_478125\|2020- | .................... | ...................A | .................... | .................... | .................... | .................... | 1 |
| D | 1222 | hCoV-19/Scotland/CVR3798/2020\|EPI_ISL_478126\|2020- | .................... | .................... | .................... | .................... | .................... | .................... | |
| D | 1223 | hCoV-19/Scotland/CVR3799/2020\|EPI_ISL_478127\|2020- | .................... | ...................A | .................... | .................... | .................... | .................... | 1 |
| D | 1224 | hCoV-19/Scotland/CVR3802/2020\|EPI_ISL_478128\|2020- | .................... | ...................A | .................... | .................... | .................... | .................... | 1 |
| D | 1225 | hCoV-19/Scotland/CVR3803/2020\|EPI_ISL_478129\|2020- | .................... | ...................A | .................... | .................... | .................... | .................... | 1 |
| D | 1226 | hCoV-19/Scotland/CVR3806/2020\|EPI_ISL_478132\|2020- | .................... | ...................A | .................... | .................... | .................... | .................... | 1 |
| D | 1228 | hCoV-19/Scotland/CVR3808/2020\|EPI_ISL_478134\|2020- | .................... | .................... | .................... | .................... | .................... | .................... | |
| D | 1229 | hCoV-19/Scotland/CVR3809/2020\|EPI_ISL_478135\|2020- | .................... | .................... | .................... | .................... | .................... | .................... | |
| D | 1230 | hCoV-19/Scotland/CVR3810/2020\|EPI_ISL_478136\|2020- | .................... | .................... | .................... | .................... | .................... | .................... | |
| D | 1231 | hCoV-19/Scotland/CVR3811/2020\|EPI_ISL_478137\|2020- | .................... | .................... | .................... | .................... | .................... | .................... | |
| D | 1232 | hCoV-19/Scotland/CVR3824/2020\|EPI_ISL_478150\|2020- | .................... | .................... | .................... | .................... | .................... | .................... | |
| D | 1233 | hCoV-19/Scotland/CVR3825/2020\|EPI_ISL_478151\|2020- | .................... | ...................A | .................... | .................... | .................... | .................... | 1 |
| D | 1234 | hCoV-19/Scotland/CVR3827/2020\|EPI_ISL_478153\|2020- | .................... | .................... | .................... | .................... | .................... | .................... | |
| D | 1235 | hCoV-19/Scotland/CVR3828/2020\|EPI_ISL_478154\|2020- | .................... | ...................A | .................... | .................... | .................... | .................... | 1 |
| D | 1236 | hCoV-19/Scotland/CVR3829/2020\|EPI_ISL_478155\|2020- | .................... | .................... | .................... | .................... | .................... | .................... | |
| D | 1237 | hCoV-19/Scotland/CVR3832/2020\|EPI_ISL_478156\|2020- | .................... | .................... | .................... | .................... | .................... | .................... | |
| D | 1238 | hCoV-19/Scotland/CVR3834/2020\|EPI_ISL_478157\|2020- | .................... | .................... | .................... | .................... | .................... | .................... | |
| D | 1239 | hCoV-19/Scotland/CVR3835/2020\|EPI_ISL_478158\|2020- | .................... | .................... | .................... | .................... | ...T................ | .................... | |
| D | 1240 | hCoV-19/Scotland/CVR3837/2020\|EPI_ISL_478159\|2020- | .................... | .................... | .................... | .................... | .................... | .................... | |
| D | 1241 | hCoV-19/Scotland/CVR3866/2020\|EPI_ISL_478160\|2020- | .................... | .................... | .................... | .................... | .................... | .................... | |
| D | 1242 | hCoV-19/Scotland/EDB4595/2020\|EPI_ISL_478165\|2020- | .................... | .................... | .................... | .................... | .................... | .................... | |
| D | 1243 | hCoV-19/Scotland/EDB4598/2020\|EPI_ISL_478166\|2020- | .T.................. | .................... | .................... | .................... | .................... | .................... | |
| D | 1244 | hCoV-19/Scotland/EDB4599/2020\|EPI_ISL_478167\|2020- | .................... | .................... | .................... | .................... | .................... | .................... | |
| D | 1245 | hCoV-19/Scotland/EDB4634/2020\|EPI_ISL_478171\|2020- | .................... | .................... | .................... | .................... | .................... | .................... | |
| D | 1246 | hCoV-19/Scotland/EDB4639/2020\|EPI_ISL_478173\|2020- | .................... | .................... | .................... | .................... | .................... | .................... | |
| D | 1247 | hCoV-19/Scotland/EDB4642/2020\|EPI_ISL_478174\|2020- | .................... | .A.................. | .................... | .................... | .................... | .................... | |
| D | 1248 | hCoV-19/Scotland/EDB4648/2020\|EPI_ISL_478176\|2020- | .................... | .................... | .................... | .................... | .................... | .................... | |
| D | 1249 | hCoV-19/Scotland/EDB4649/2020\|EPI_ISL_478177\|2020- | .................... | .................... | .................... | .................... | .................... | .................... | |
| D | 1250 | hCoV-19/Scotland/EDB4651/2020\|EPI_ISL_478178\|2020- | .................... | .................... | .................... | .................... | .................... | .................... | |
| D | 1251 | hCoV-19/Scotland/EDB4652/2020\|EPI_ISL_478179\|2020- | .................... | .................... | .................... | .................... | .................... | .................... | |
| D | 1252 | hCoV-19/Wales/PHWC-16519C/2020\|EPI_ISL_479431\|2020 | .................... | .................... | .................... | .................... | .................... | .................... | |
| D | 1253 | hCoV-19/Wales/PHWC-165220/2020\|EPI_ISL_479436\|2020 | .................... | .................... | .................... | .................... | .................... | .................... | |
| D | 1254 | hCoV-19/Wales/PHWC-16525D/2020\|EPI_ISL_479439\|2020 | .................... | .................... | .................... | .................... | .................... | .................... | |
| D | 1255 | hCoV-19/Wales/PHWC-16527B/2020\|EPI_ISL_479441\|2020 | .................... | .................... | .................... | .................... | .................... | .................... | |
| D | 1256 | hCoV-19/Wales/PHWC-16528A/2020\|EPI_ISL_479442\|2020 | .................... | .................... | .................... | .................... | .................... | .................... | |
| D | 1257 | hCoV-19/Wales/PHWC-165299/2020\|EPI_ISL_479443\|2020 | .................... | .................... | .................... | .................... | .................... | .................... | |
| D | 1258 | hCoV-19/Wales/PHWC-1652B7/2020\|EPI_ISL_479445\|2020 | .................... | .................... | .................... | .................... | .................... | .................... | |

| | | | |
|---|---|---|---|
| D | 1259 | hCoV-19/Wales/PHWC-16531E/2020\|EPI_ISL_479451\|2020 | .................... .................... .................... .................... .................... .................... |
| D | 1260 | hCoV-19/Wales/PHWC-165378/2020\|EPI_ISL_479455\|2020 | .................... .................... .................... .................... .................... .................... |
| D | 1261 | hCoV-19/Wales/PHWC-16542A/2020\|EPI_ISL_479463\|2020 | .................... .................... .................... .................... .................... .................... |
| D | 1262 | hCoV-19/Wales/PHWC-165581/2020\|EPI_ISL_479480\|2020 | .................... .................... .................... .................... .................... .................... |
| D | 1263 | hCoV-19/Singapore/526/2020\|EPI_ISL_479482\|2020-04- | .................... .................... .................... .................... .................... .................... |
| D | 1264 | hCoV-19/Singapore/527/2020\|EPI_ISL_479483\|2020-04- | .................... .................... .................... .................... .................... .................... |
| D | 1265 | hCoV-19/Singapore/528/2020\|EPI_ISL_479484\|2020-04- | .................... .................... .................... .................... .................... .................... |
| D | 1266 | hCoV-19/Singapore/529/2020\|EPI_ISL_479485\|2020-04- | .................... .................... .................... .................... .................... .................... |
| D | 1267 | hCoV-19/Singapore/530/2020\|EPI_ISL_479486\|2020-04- | .................... .................... .................... .................... .................... .................... |
| D | 1268 | hCoV-19/Singapore/531/2020\|EPI_ISL_479487\|2020-04- | .................... .................... .................... .................... .................... .................... |
| D | 1269 | hCoV-19/Singapore/532/2020\|EPI_ISL_479488\|2020-04- | .................... .................... .................... .................... .................... .................... |
| D | 1270 | hCoV-19/Singapore/533/2020\|EPI_ISL_479489\|2020-04- | .................... .................... .................... .................... .................... .................... |
| D | 1271 | hCoV-19/Singapore/534/2020\|EPI_ISL_479490\|2020-04- | .................... .................... .................... .................... .................... .................... |
| D | 1272 | hCoV-19/Singapore/535/2020\|EPI_ISL_479491\|2020-04- | .................... .................... .................... .................... .................... .................... |
| D | 1273 | hCoV-19/Singapore/536/2020\|EPI_ISL_479492\|2020-04- | .................... .................... .................... .................... .................... .................... |
| D | 1274 | hCoV-19/India/MH-NIV-10176/2020\|EPI_ISL_479493\|202 | .................... .................... .................... .................... .................... .................... |
| D | 1275 | hCoV-19/India/MH-NIV-11670/2020\|EPI_ISL_479494\|202 | .................... .................... .................... .................... .................... .................... |
| D | 1276 | hCoV-19/India/MH-NIV-11857/2020\|EPI_ISL_479495\|202 | .................... .................... .................... .................... .................... .................... |
| D | 1277 | hCoV-19/India/MH-NIV-12430/2020\|EPI_ISL_479496\|202 | .................... .................... .................... .................... .................... .................... |
| D | 1278 | hCoV-19/India/MH-NIV-12628/2020\|EPI_ISL_479497\|202 | .................... .................... .................... .................... .................... .................... |
| D | 1280 | hCoV-19/India/MH-NIV-14614/2020\|EPI_ISL_479499\|202 | .................... .................... .................... .................... .................... .................... |
| D | 1281 | hCoV-19/Singapore/192/2020\|EPI_ISL_462310\|2020-04- | .................... .................... .................... .................... .................... .................... |
| D | 1282 | hCoV-19/Singapore/193/2020\|EPI_ISL_462311\|2020-04- | .................... .................... .................... .................... .................... .................... |
| D | 1283 | hCoV-19/Singapore/194/2020\|EPI_ISL_462312\|2020-04- | .................... .................... .................... .................... .................... .................... |
| D | 1284 | hCoV-19/Singapore/195/2020\|EPI_ISL_462313\|2020-04- | .................... .................... .................... .................... .................... .................... |
| D | 1285 | hCoV-19/Singapore/196/2020\|EPI_ISL_462314\|2020-04- | .................... .................... .................... .................... .................... .................... |
| D | 1286 | hCoV-19/Singapore/197/2020\|EPI_ISL_462315\|2020-04- | .................... .................... .................... .................... .................... .................... |
| D | 1287 | hCoV-19/Singapore/198/2020\|EPI_ISL_462316\|2020-04- | .................... .................... .................... .................... .................... .................... |
| D | 1288 | hCoV-19/Singapore/199/2020\|EPI_ISL_462317\|2020-04- | .................... .................... .................... .................... .................... .................... |
| D | 1289 | hCoV-19/Singapore/200/2020\|EPI_ISL_462318\|2020-04- | .................... .................... .................... .................... .................... .................... |
| D | 1290 | hCoV-19/Singapore/201/2020\|EPI_ISL_462319\|2020-04- | .................... .................... .................... .................... .................... .................... |
| D | 1291 | hCoV-19/Singapore/202/2020\|EPI_ISL_462320\|2020-04- | .................... .................... .................... .................... .................... .................... |
| D | 1292 | hCoV-19/Singapore/203/2020\|EPI_ISL_462321\|2020-04- | .................... .................... .................... .................... .................... .................... |
| D | 1293 | hCoV-19/Singapore/204/2020\|EPI_ISL_462322\|2020-04- | .................... .................... .................... .................... .................... .................... |
| D | 1294 | hCoV-19/Singapore/205/2020\|EPI_ISL_462323\|2020-04- | .................... .................... .................... .................... .................... .................... |
| D | 1295 | hCoV-19/Singapore/206/2020\|EPI_ISL_462324\|2020-04- | .................... .................... .................... .................... .................... .................... |
| D | 1296 | hCoV-19/Singapore/207/2020\|EPI_ISL_462325\|2020-04- | .................... .................... .................... .................... .................... .................... |
| D | 1297 | hCoV-19/Singapore/208/2020\|EPI_ISL_462326\|2020-04- | .................... .................... .................... .................... .................... .................... |
| D | 1298 | hCoV-19/Singapore/210/2020\|EPI_ISL_462328\|2020-03- | .................... .................... .................... .................... .................... .................... |
| D | 1299 | hCoV-19/Singapore/220/2020\|EPI_ISL_462338\|2020-03- | .................... .................... .................... .................... .................... .................... |
| D | 1300 | hCoV-19/Singapore/222/2020\|EPI_ISL_462340\|2020-04- | .................... .................... .................... .................... .................... .................... |
| D | 1301 | hCoV-19/Singapore/223/2020\|EPI_ISL_462341\|2020-03- | .................... .................... .................... .................... .................... .................... |
| D | 1302 | hCoV-19/Singapore/224/2020\|EPI_ISL_462342\|2020-03- | .................... .................... .................... .................... .................... .................... |
| D | 1303 | hCoV-19/Singapore/225/2020\|EPI_ISL_462343\|2020-04- | .................... .................... .................... .................... .................... .................... |
| D | 1304 | hCoV-19/Singapore/226/2020\|EPI_ISL_462344\|2020-04- | .................... .................... .................... .................... .................... .................... |
| D | 1305 | hCoV-19/Singapore/227/2020\|EPI_ISL_462345\|2020-03- | .................... .................... .................... .................... .................... .................... |
| D | 1306 | hCoV-19/Singapore/228/2020\|EPI_ISL_462346\|2020-03- | .................... .................... .................... .................... .................... .................... |
| D | 1307 | hCoV-19/Singapore/229/2020\|EPI_ISL_462347\|2020-04- | .................... .................... .................... .................... .................... .................... |
| D | 1308 | hCoV-19/Singapore/230/2020\|EPI_ISL_462348\|2020-04- | .................... .................... .................... .................... .................... .................... |
| D | 1309 | hCoV-19/Singapore/231/2020\|EPI_ISL_462349\|2020-03- | .................... .................... .................... .................... .................... .................... |
| D | 1310 | hCoV-19/Singapore/232/2020\|EPI_ISL_462350\|2020-03- | .................... .................... .................... .................... .................... .................... |
| D | 1311 | hCoV-19/Singapore/233/2020\|EPI_ISL_462351\|2020-03- | .................... .................... .................... .................... .................... .................... |
| D | 1312 | hCoV-19/Singapore/234/2020\|EPI_ISL_462352\|2020-04- | .................... .................... .................... .................... .................... .................... |
| D | 1313 | hCoV-19/Singapore/235/2020\|EPI_ISL_462353\|2020-04- | .................... .................... .................... .................... .................... .................... |
| D | 1314 | hCoV-19/Singapore/258/2020\|EPI_ISL_462376\|2020-03- | .................... .................... .................... .................... .................... .................... |
| D | 1315 | hCoV-19/Singapore/259/2020\|EPI_ISL_462377\|2020-04- | .................... .................... .................... .................... .................... .................... |
| D | 1316 | hCoV-19/Singapore/260/2020\|EPI_ISL_462378\|2020-04- | .................... .................... .................... .................... .................... .................... |
| D | 1317 | hCoV-19/Singapore/261/2020\|EPI_ISL_462379\|2020-04- | .................... .................... .................... .................... .................... .................... |
| D | 1318 | hCoV-19/Singapore/262/2020\|EPI_ISL_462380\|2020-04- | .................... .................... .................... .................... .................... .................... |
| D | 1319 | hCoV-19/Singapore/263/2020\|EPI_ISL_462381\|2020-04- | .................... .................... .................... .................... .................... .................... |
| D | 1320 | hCoV-19/Singapore/264/2020\|EPI_ISL_462382\|2020-05- | .................... .................... .................... .................... .................... .................... |
| D | 1321 | hCoV-19/Singapore/265/2020\|EPI_ISL_462383\|2020-04- | .................... .................... .................... .................... .................... .................... |
| D | 1322 | hCoV-19/Singapore/266/2020\|EPI_ISL_462384\|2020-04- | .................... .................... .................... .................... .................... .................... |
| D | 1323 | hCoV-19/Singapore/267/2020\|EPI_ISL_462385\|2020-03- | .................... .................... .................... .................... .................... .................... |
| D | 1324 | hCoV-19/Singapore/268/2020\|EPI_ISL_462386\|2020-03- | .................... .................... .................... .................... .................... .................... |
| D | 1325 | hCoV-19/Singapore/269/2020\|EPI_ISL_462387\|2020-03- | .................... .................... .................... .................... .................... .................... |
| D | 1326 | hCoV-19/Singapore/270/2020\|EPI_ISL_462388\|2020-04- | .................... .................... .................... .................... .................... .................... |
| D | 1327 | hCoV-19/Singapore/271/2020\|EPI_ISL_462389\|2020-03- | .................... .................... .................... .................... .................... .................... |
| D | 1328 | hCoV-19/Singapore/272/2020\|EPI_ISL_462390\|2020-03- | .................... .................... .................... .................... .................... .................... |
| D | 1329 | hCoV-19/Singapore/273/2020\|EPI_ISL_462391\|2020-03- | .................... .................... .................... .................... .................... .................... |
| D | 1330 | hCoV-19/Singapore/274/2020\|EPI_ISL_462392\|2020-03- | .................... .................... .................... .................... .................... .................... |
| D | 1331 | hCoV-19/Singapore/275/2020\|EPI_ISL_462393\|2020-05- | .................... .................... .................... .................... .................... .................... |
| D | 1332 | hCoV-19/Singapore/276/2020\|EPI_ISL_462394\|2020-04- | .................... .................... .................... .................... .................... .................... |
| D | 1333 | hCoV-19/Singapore/277/2020\|EPI_ISL_462395\|2020-04- | .................... .................... .................... .................... .................... .................... |
| D | 1334 | hCoV-19/Singapore/278/2020\|EPI_ISL_462396\|2020-03- | .................... .................... .................... .................... .................... .................... |
| D | 1335 | hCoV-19/Singapore/279/2020\|EPI_ISL_462397\|2020-04- | .................... .................... .................... .................... .................... .................... |
| D | 1336 | hCoV-19/USA/WA-S1151/2020\|EPI_ISL_463641\|2020-05-0 | .................... .................... .................... .................... .................... .................... |
| D | 1337 | hCoV-19/USA/WA-S1152/2020\|EPI_ISL_463642\|2020-05-0 | .................... .................... .................... .................... .................... .................... |

```
D 1338 hCoV-19/USA/WA-S1153/2020|EPI_ISL_463643|2020-05-0 ···················· ···················· ···················· ···················· ···················· ····················
D 1339 hCoV-19/USA/WA-S1154/2020|EPI_ISL_463644|2020-05-0 ···················· ···················· ···················· ···················· ···················· ····················
D 1340 hCoV-19/USA/WA-S1155/2020|EPI_ISL_463645|2020-05-0 ···················· ···················· ···················· ···················· ···················· ····················
D 1341 hCoV-19/USA/WA-S1156/2020|EPI_ISL_463646|2020-05-0 ···················· ···················· ···················· ···················· ···················· ····················
D 1342 hCoV-19/USA/WA-S1157/2020|EPI_ISL_463647|2020-05-0 ···················· ···················· ···················· ···················· ···················· ····················
D 1343 hCoV-19/USA/WA-S1158/2020|EPI_ISL_463648|2020-05-0 ···················· ···················· ···················· ···················· ···················· ····················
D 1344 hCoV-19/USA/WA-S1159/2020|EPI_ISL_463649|2020-05-0 ···················· ···················· ···················· ···················· ···················· ····················
D 1345 hCoV-19/USA/WA-S1160/2020|EPI_ISL_463650|2020-05-0 ···················· ···················· ···················· ···················· ···················· ····················
D 1346 hCoV-19/USA/WA-S1161/2020|EPI_ISL_463651|2020-05-0 ···················· ···················· ···················· ···················· ···················· ····················
D 1347 hCoV-19/USA/WA-S1164/2020|EPI_ISL_463654|2020-05-0 ···················· ···················· ···················· ···················· ···················· ····················
D 1348 hCoV-19/USA/WA-S1165/2020|EPI_ISL_463655|2020-05-0 ···················· ···················· ···················· ···················· ···················· ····················
D 1349 hCoV-19/USA/WA-S1166/2020|EPI_ISL_463656|2020-05-0 ···················· ···················· ···················· ···················· ···················· ····················
D 1350 hCoV-19/USA/WA-S1167/2020|EPI_ISL_463657|2020-05-0 ···················· ···················· ···················· ···················· ···················· ····················
D 1351 hCoV-19/USA/WA-S1168/2020|EPI_ISL_463658|2020-05-0 ···················· ···················· ···················· ···················· ···················· ····················
D 1352 hCoV-19/USA/WA-S1169/2020|EPI_ISL_463659|2020-05-0 ···················· ···················· ···················· ···················· ···················· ····················
D 1353 hCoV-19/USA/WA-S1170/2020|EPI_ISL_463660|2020-05-0 ···················· ···················· ···················· ···················· ···················· ····················
D 1354 hCoV-19/USA/WA-S1171/2020|EPI_ISL_463661|2020-05-0 ···················· ···················· ···················· ···················· ···················· ····················
D 1355 hCoV-19/USA/WA-S1172/2020|EPI_ISL_463662|2020-04-2 ···················· ···················· ···················· ···················· ···················· ····················
D 1356 hCoV-19/USA/WA-S1173/2020|EPI_ISL_463663|2020-05-0 ···················· ···················· ···················· ···················· ···················· ····················
D 1357 hCoV-19/USA/WA-S1174/2020|EPI_ISL_463664|2020-05-0 ···················· ···················· ···················· ···················· ···················· ····················
D 1358 hCoV-19/USA/WA-S1175/2020|EPI_ISL_463665|2020-05-0 ···················· ·A·················· ···················· ···················· ···················· ····················
D 1359 hCoV-19/USA/WA-S1176/2020|EPI_ISL_463666|2020-05-0 ···················· ···················· ···················· ···················· ···················· ····················
D 1360 hCoV-19/USA/WA-S1177/2020|EPI_ISL_463667|2020-05-0 ···················· ···················· ···················· ···················· ···················· ····················
D 1361 hCoV-19/USA/WA-S1178/2020|EPI_ISL_463668|2020-05-0 ···················· ···················· ···················· ···················· ···················· ····················
D 1362 hCoV-19/USA/WA-S1179/2020|EPI_ISL_463669|2020-05-0 ···················· ···················· ···················· ···················· ···················· ····················
D 1363 hCoV-19/USA/WA-S1180/2020|EPI_ISL_463670|2020-05-0 ···················· ···················· ···················· ···················· ···················· ····················
D 1364 hCoV-19/USA/WA-S1181/2020|EPI_ISL_463671|2020-05-0 ···················· ···················· ···················· ···················· ···················· ····················
D 1365 hCoV-19/USA/WA-S1182/2020|EPI_ISL_463672|2020-05-0 ···················· ···················· ···················· ···················· ···················· ····················
D 1366 hCoV-19/USA/WA-S1183/2020|EPI_ISL_463673|2020-05-0 ···················· ···················· ···················· ···················· ···················· ····················
D 1367 hCoV-19/USA/WA-S1184/2020|EPI_ISL_463674|2020-05-0 ···················· ···················· ···················· ···················· ···················· ····················
D 1368 hCoV-19/USA/WA-S1185/2020|EPI_ISL_463675|2020-05-0 ···················· ···················· ···················· ···················· ···················· ····················
D 1369 hCoV-19/USA/WA-S1186/2020|EPI_ISL_463676|2020-05-0 ···················· ···················· ···················· ···················· ···················· ····················
D 1370 hCoV-19/USA/WA-S1187/2020|EPI_ISL_463677|2020-05-0 ···················· ···················· ···················· ···················· ···················· ····················
D 1371 hCoV-19/USA/WA-S1188/2020|EPI_ISL_463678|2020-05-0 ···················· ···················· ···················· ···················· ···················· ····················
D 1372 hCoV-19/USA/WA-S1189/2020|EPI_ISL_463679|2020-05-0 ···················· ···················· ···················· ···················· ···················· ····················
D 1373 hCoV-19/USA/WA-S1190/2020|EPI_ISL_463680|2020-05-0 ···················· ···················· ···················· ···················· ···················· ····················
D 1374 hCoV-19/USA/WA-S1191/2020|EPI_ISL_463681|2020-05-0 ···················· ···················· ···················· ···················· ···················· ····················
D 1375 hCoV-19/USA/WA-S1192/2020|EPI_ISL_463682|2020-05-0 ···················· ·A·················· ···················· ···················· ···················· ····················
D 1376 hCoV-19/USA/WA-S1194/2020|EPI_ISL_463684|2020-05-0 ···················· ···················· ···················· ···················· ···················· ····················
D 1378 hCoV-19/England/20118143704/2020|EPI_ISL_464973|20 ···················· ···················· ···················· ···················· ···················· ····················
D 1379 hCoV-19/England/20118146504/2020|EPI_ISL_464974|20 ···················· ···················· ···················· ···················· ···················· ····················
D 1380 hCoV-19/England/20118147904/2020|EPI_ISL_464976|20 ···················· ···················· ···················· ···················· ···················· ····················
D 1381 hCoV-19/England/20118148804/2020|EPI_ISL_464978|20 ···················· ···················· ···················· ···················· ···················· ····················
D 1382 hCoV-19/England/20118151104/2020|EPI_ISL_464979|20 ···················· ···················· ···················· ···················· ··N················· ····················
D 1383 hCoV-19/England/20118151404/2020|EPI_ISL_464980|20 ···················· ···················· ···················· ···················· ···················· ····················
D 1384 hCoV-19/England/20118158504/2020|EPI_ISL_464985|20 ···················· ···················· ···················· ···················· ···················· ····················
D 1385 hCoV-19/England/20118164704/2020|EPI_ISL_464989|20 ···················· ···················· ···················· ···················· ···················· ····················
D 1386 hCoV-19/England/20118171704/2020|EPI_ISL_464991|20 ···················· ···················· ···················· ···················· ···················· ····················
D 1387 hCoV-19/England/20118175304/2020|EPI_ISL_464996|20 ···················· ···················· ···················· ···················· ···················· ····················
D 1388 hCoV-19/England/20118176104/2020|EPI_ISL_464997|20 ···················· ···················· ···················· ···················· ···················· ····················
D 1389 hCoV-19/England/20118178504/2020|EPI_ISL_464999|20 ···················· ···················· ···················· ···················· ···················· ····················
D 1390 hCoV-19/USA/CA-CZB-1563/2020|EPI_ISL_468500|2020-0 ···················· ···················· ···················· ···················· ···················· ····················
D 1391 hCoV-19/USA/CA-CZB-1564/2020|EPI_ISL_468501|2020-0 ···················· ···················· ···················· ···················· ···················· ····················
D 1392 hCoV-19/USA/CA-CZB-1566/2020|EPI_ISL_468502|2020-0 ···················· ···················· ···················· ···················· ···················· ····················
D 1393 hCoV-19/USA/CA-CZB-1570/2020|EPI_ISL_468503|2020-0 ···················· ···················· ···················· ···················· ···················· ····················
D 1394 hCoV-19/USA/CA-CZB-1572/2020|EPI_ISL_468504|2020-0 ···················· ···················· ···················· ···················· ···················· ····················
D 1395 hCoV-19/USA/CA-CZB-1573/2020|EPI_ISL_468505|2020-0 ···················· ···················· ···················· ···················· ···················· ····················
D 1396 hCoV-19/USA/CA-CZB-1575/2020|EPI_ISL_468506|2020-0 ···················· ···················· ···················· ···················· N··················· ····················
D 1397 hCoV-19/USA/CA-CZB-1576/2020|EPI_ISL_468507|2020-0 ···················· ···················· ···················· ···················· ···················· ····················
D 1398 hCoV-19/USA/CA-CZB-1577/2020|EPI_ISL_468508|2020-0 ···················· ···················· ···················· ···················· ···················· ····················
D 1399 hCoV-19/USA/CA-CZB-1578/2020|EPI_ISL_468509|2020-0 ···················· ···················· ···················· ···················· ···················· ····················
D 1400 hCoV-19/USA/CA-CZB-1579/2020|EPI_ISL_468510|2020-0 ···················· ···················· ···················· ···················· ···················· ····················
D 1401 hCoV-19/USA/CA-CZB-1581/2020|EPI_ISL_468511|2020-0 ···················· ···················· ···················· ···················· ···················· ····················
D 1402 hCoV-19/USA/CA-CZB-1582/2020|EPI_ISL_468512|2020-0 ···················· ···················· ···················· ···················· ···················· ····················
D 1403 hCoV-19/USA/CA-CZB-1583/2020|EPI_ISL_468513|2020-0 ···················· ···················· ···················· ···················· ···················· ····················
D 1404 hCoV-19/England/BRIS-12B165/2020|EPI_ISL_469827|20 ···················· ···················· ···················· ···················· ···················· ····················
D 1405 hCoV-19/England/BRIS-12B1A1/2020|EPI_ISL_469829|20 ···················· ···················· ···················· ···················· ···················· ····················
D 1406 hCoV-19/England/BRIS-12B0FF/2020|EPI_ISL_469831|20 ···················· ···················· ···················· ···················· ···················· ····················
D 1407 hCoV-19/England/BRIS-12B1CF/2020|EPI_ISL_469833|20 ···················· ···················· ···················· ···················· ···················· ····················
D 1408 hCoV-19/England/BRIS-12B208/2020|EPI_ISL_469837|20 ···················· ···················· ···················· ···················· ···················· ····················
D 1409 hCoV-19/England/BRIS-12B068/2020|EPI_ISL_469841|20 ···················· ···················· ···················· ···················· ···················· ····················
D 1410 hCoV-19/USA/MN-MDH-1120/2020|EPI_ISL_470800|2020-0 ···················· ·A·················· ···················· ···················· ···················· ····················
D 1411 hCoV-19/Bangladesh/icddrb-2083/2020|EPI_ISL_470801 ···················· ···················· ···················· ···················· ···················· ····················
D 1412 hCoV-19/Wales/PHWC-1609E4/2020|EPI_ISL_473002|2020 ···················· ···················· ···················· ···················· ···················· ····················
D 1413 hCoV-19/Wales/PHWC-160A1E/2020|EPI_ISL_473004|2020 ···················· ···················· ···················· ···················· ···················· ····················
D 1414 hCoV-19/Wales/PHWC-160A3C/2020|EPI_ISL_473005|2020 ···················· ···················· ···················· ···················· ···················· ····················
D 1415 hCoV-19/Wales/PHWC-160AA5/2020|EPI_ISL_473009|2020 ···················· ···················· ···················· ···················· ···················· ····················
D 1416 hCoV-19/Wales/PHWC-160AB4/2020|EPI_ISL_473010|2020 ···················· ···················· ···················· ···················· ···················· ····················
```

D 1417 hCoV-19/Wales/PHWC-160B0C/2020|EPI_ISL_473014|2020 ···················· ···················· ····················· ···················· ···················· ····················
D 1418 hCoV-19/Wales/PHWC-160B1B/2020|EPI_ISL_473015|2020 ···················· ···················· ····················· ···················· ···················· ····················
D 1419 hCoV-19/Wales/PHWC-160B57/2020|EPI_ISL_473019|2020 ···················· ···················· ····················· ···················· ···················· ····················
D 1420 hCoV-19/Wales/PHWC-160B66/2020|EPI_ISL_473020|2020 ···················· ···················· ····················· ···················· ···················· ····················
D 1421 hCoV-19/Wales/PHWC-160B75/2020|EPI_ISL_473021|2020 ···················· ···················· ····················· ···················· ···················· ····················
D 1422 hCoV-19/Wales/PHWC-160BA2/2020|EPI_ISL_473024|2020 ···················· ···················· ····················· ···················· ···················· ····················
D 1423 hCoV-19/Wales/PHWC-160BC0/2020|EPI_ISL_473026|2020 ···················· ···················· ····················· ···················· ···················· ····················
D 1424 hCoV-19/Wales/PHWC-160BFD/2020|EPI_ISL_473027|2020 ···················· ···················· ····················· ···················· ···················· ····················
D 1425 hCoV-19/Wales/PHWC-160C18/2020|EPI_ISL_473029|2020 ···················· ···················· ····················· ···················· ···················· ····················
D 1426 hCoV-19/Wales/PHWC-160C36/2020|EPI_ISL_473031|2020 ···················· ···················· ····················· ···················· ···················· ····················
D 1427 hCoV-19/Wales/PHWC-160C54/2020|EPI_ISL_473032|2020 ···················· ····A··············· ····················· ···················· ···················· ····················
D 1428 hCoV-19/Wales/PHWC-160C63/2020|EPI_ISL_473033|2020 ···················· ···················· ····················· ···················· ···················· ····················
D 1429 hCoV-19/Wales/PHWC-160C81/2020|EPI_ISL_473034|2020 ···················· ···················· ····················· ···················· ···················· ····················
D 1430 hCoV-19/Wales/PHWC-160C90/2020|EPI_ISL_473035|2020 ···················· ···················· ····················· ···················· T··················· ····················
D 1431 hCoV-19/Wales/PHWC-160CAF/2020|EPI_ISL_473036|2020 ···················· ···················· ····················· ···················· ···················· ····················
D 1432 hCoV-19/Wales/PHWC-160CBE/2020|EPI_ISL_473037|2020 ···················· ···················· ····················· ···················· ···················· ····················
D 1433 hCoV-19/Wales/PHWC-160CCD/2020|EPI_ISL_473038|2020 ···················· ···················· ····················· ···················· ···················· ····················
D 1434 hCoV-19/Wales/PHWC-160CDC/2020|EPI_ISL_473039|2020 ···················· ···················· ····················· ···················· ···················· ····················
D 1435 hCoV-19/Wales/PHWC-160D33/2020|EPI_ISL_473044|2020 ···················· ···················· ····················· ···················· ···················· ····················
D 1436 hCoV-19/Wales/PHWC-160D42/2020|EPI_ISL_473045|2020 ···················· ···················· ····················· ···················· ···················· ····················
D 1437 hCoV-19/Wales/PHWC-16244B/2020|EPI_ISL_473074|2020 ···················· ···················· ····················· ···················· ···················· ····················
D 1438 hCoV-19/Wales/PHWC-1624D2/2020|EPI_ISL_473079|2020 ···················· ···················· ····················· ···················· ···················· ····················
D 1439 hCoV-19/Wales/PHWC-1624F0/2020|EPI_ISL_473081|2020 ···················· ···················· ····················· ···················· ···················· ····················
D 1440 hCoV-19/Wales/PHWC-162539/2020|EPI_ISL_473085|2020 ···················· ···················· ····················· ···················· ···················· ····················
D 1441 hCoV-19/Wales/PHWC-162566/2020|EPI_ISL_473088|2020 ···················· ···················· ····················· ···················· ···················· ····················
D 1442 hCoV-19/Wales/PHWC-2C29F/2020|EPI_ISL_474301|2020- ···················· ···················· ····················· ···················· ···················· ····················
D 1443 hCoV-19/Wales/PHWC-2C2AE/2020|EPI_ISL_474302|2020- ···················· ···················· ····················· ···················· ···················· ····················
D 1444 hCoV-19/Wales/PHWC-2C2BD/2020|EPI_ISL_474303|2020- ···················· ···················· ····················· ···················· ···················· ····················
D 1445 hCoV-19/Wales/PHWC-325F7/2020|EPI_ISL_474333|2020- ···················· ···················· ····················· ···················· ···················· ····················
D 1446 hCoV-19/Wales/PHWC-32621/2020|EPI_ISL_474336|2020- ···················· ···················· ····················· ···················· ···················· ····················
D 1447 hCoV-19/Wales/PHWC-326B8/2020|EPI_ISL_474340|2020- ···················· ···················· ····················· ···················· ···················· ····················
D 1448 hCoV-19/Wales/PHWC-32700/2020|EPI_ISL_474343|2020- ···················· ···················· ····················· ···················· ···················· ····················
D 1449 hCoV-19/Wales/PHWC-32779/2020|EPI_ISL_474349|2020- ···················· ···················· ····················· ···················· ···················· ····················
D 1450 hCoV-19/Wales/PHWC-327F1/2020|EPI_ISL_474352|2020- ···················· ····A··············· ····················· ···················· ···················· ····················
D 1451 hCoV-19/Wales/PHWC-3280D/2020|EPI_ISL_474353|2020- ···················· ···················· ····················· ···················· ···················· ····················
D 1452 hCoV-19/Wales/PHWC-3282B/2020|EPI_ISL_474355|2020- ···················· ···················· ····················· ···················· ···················· ····················
D 1453 hCoV-19/Wales/PHWC-3283A/2020|EPI_ISL_474356|2020- ···················· ···················· ····················· ···················· ···················· ····················
D 1454 hCoV-19/Wales/PHWC-32858/2020|EPI_ISL_474358|2020- ···················· ···················· ····················· ···················· ···················· ····················
D 1455 hCoV-19/Wales/PHWC-32876/2020|EPI_ISL_474359|2020- ···················· ···················· ····················· ···················· ···················· ····················
D 1456 hCoV-19/Wales/PHWC-32885/2020|EPI_ISL_474360|2020- ···················· ····A··············· ····················· ···················· ···················· ····················
D 1457 hCoV-19/Wales/PHWC-328B2/2020|EPI_ISL_474361|2020- ···················· ···················· ····················· ···················· ···················· ····················
D 1458 hCoV-19/Wales/PHWC-328C1/2020|EPI_ISL_474362|2020- ···················· ···················· ····················· ···················· ···················· ····················
D 1459 hCoV-19/Wales/PHWC-328EF/2020|EPI_ISL_474364|2020- ···················· ···················· ····················· ···················· ···················· ····················
D 1460 hCoV-19/Wales/PHWC-32919/2020|EPI_ISL_474367|2020- ···················· ···················· ····················· ···················· ···················· ····················
D 1461 hCoV-19/Wales/PHWC-32928/2020|EPI_ISL_474368|2020- ···················· ····A··············· ····················· ···················· ···················· ····················
D 1462 hCoV-19/Wales/PHWC-32937/2020|EPI_ISL_474369|2020- ···················· ···················· ····················· ···················· ···················· ····················
D 1463 hCoV-19/Wales/PHWC-32955/2020|EPI_ISL_474370|2020- ···················· ···················· ····················· ···················· ···················· ····················
D 1464 hCoV-19/Wales/PHWC-329A0/2020|EPI_ISL_474375|2020- ···················· ···················· ····················· ···················· ···················· ····················
D 1465 hCoV-19/Wales/PHWC-329BF/2020|EPI_ISL_474376|2020- ···················· ···················· ····················· ···················· ···················· ····················
D 1466 hCoV-19/USA/CA-CSMC142/2020|EPI_ISL_475621|2020-04 ···················· ···················· ····················· ···················· ···················· ····················
D 1467 hCoV-19/USA/CA-CSMC143/2020|EPI_ISL_475622|2020-04 ···················· ···················· ····················· ···················· ···················· ····················
D 1468 hCoV-19/USA/CA-CSMC16/2020|EPI_ISL_475625|2020-04- ···················· ···················· ····················· ···················· ···················· ····················
D 1469 hCoV-19/USA/CA-CSMC18/2020|EPI_ISL_475627|2020-03- ···················· ···················· ····················· ···················· ···················· ····················
D 1470 hCoV-19/USA/CA-CSMC19/2020|EPI_ISL_475628|2020-03- ···················· ···················· ····················· ···················· ···················· ····················
D 1471 hCoV-19/USA/CA-CSMC2/2020|EPI_ISL_475629|2020-03-2 ···················· ···················· ····················· ···················· ···················· ····················
D 1472 hCoV-19/USA/CA-CSMC20/2020|EPI_ISL_475630|2020-03- ···················· ···················· ····················· ···················· ···················· ····················
D 1473 hCoV-19/USA/CA-CSMC21/2020|EPI_ISL_475631|2020-03- ···················· ···················· ····················· ···················· ···················· ····················
D 1474 hCoV-19/USA/CA-CSMC22/2020|EPI_ISL_475632|2020-03- ···················· ···················· ····················· ···················· ···················· ····················
D 1475 hCoV-19/USA/CA-CSMC23/2020|EPI_ISL_475633|2020-03- ···················· ···················· ····················· ···················· ···················· ····················
D 1476 hCoV-19/USA/CA-CSMC24/2020|EPI_ISL_475634|2020-03- ···················· ···················· ····················· ···················· ···················· ····················
D 1477 hCoV-19/USA/CA-CSMC25/2020|EPI_ISL_475635|2020-03- ···················· ···················· ····················· ···················· ···················· ····················
D 1478 hCoV-19/USA/CA-CSMC26/2020|EPI_ISL_475636|2020-03- ···················· ···················· ····················· ···················· ···················· ····················
D 1479 hCoV-19/USA/CA-CSMC27/2020|EPI_ISL_475637|2020-03- ···················· ···················· ····················· ···················· ···················· ····················
D 1480 hCoV-19/USA/CA-CSMC28/2020|EPI_ISL_475638|2020-03- ···················· ···················· ····················· ···················· ···················· ····················
D 1481 hCoV-19/USA/CA-CSMC29/2020|EPI_ISL_475639|2020-03- ···················· ···················· ····················· ···················· ···················· ····················
D 1482 hCoV-19/USA/CA-CSMC51/2020|EPI_ISL_475664|2020-04- ···················· ···················· ····················· ···················· ···················· ····················
D 1483 hCoV-19/USA/CA-CSMC53/2020|EPI_ISL_475666|2020-04- ···················· ···················· ····················· ···················· ···················· ····················
D 1484 hCoV-19/USA/CA-CSMC54/2020|EPI_ISL_475667|2020-04- ···················· ···················· ····················· ···················· ···················· ····················
D 1485 hCoV-19/USA/CA-CSMC55/2020|EPI_ISL_475668|2020-04- ···················· ···················· ····················· ···················· ···················· ····················
D 1486 hCoV-19/USA/CA-CSMC56/2020|EPI_ISL_475669|2020-04- ···················· ···················· ····················· ···················· ···················· ····················
D 1487 hCoV-19/USA/CA-CSMC57/2020|EPI_ISL_475670|2020-04- ···················· ···················· ····················· ···················· ···················· ····················
D 1488 hCoV-19/USA/CA-CSMC58/2020|EPI_ISL_475671|2020-04- ···················· ···················· ····················· ···················· ···················· ····················
D 1489 hCoV-19/USA/CA-CSMC6/2020|EPI_ISL_475673|2020-03-2 ···················· ···················· ····················· ···················· ···················· ····················
D 1490 hCoV-19/USA/CA-CSMC60/2020|EPI_ISL_475674|2020-04- ···················· ···················· ····················· ···················· ···················· ····················
D 1491 hCoV-19/USA/CA-CSMC61/2020|EPI_ISL_475675|2020-04- ···················· ···················· ····················· ···················· ···················· ····················
D 1492 hCoV-19/USA/CA-CSMC63/2020|EPI_ISL_475677|2020-03- ···················· ···················· ····················· ···················· ···················· ····················
D 1493 hCoV-19/USA/CA-CSMC64/2020|EPI_ISL_475678|2020-03- ···················· ···················· ····················· ···················· ···················· ····················
D 1494 hCoV-19/USA/CA-CSMC65/2020|EPI_ISL_475679|2020-03- ···················· ···················· ····················· ···················· ···················· ····················

| | | | | | | | | |
|---|---|---|---|---|---|---|---|---|
| D | 1495 | hCoV-19/USA/CA-CSMC66/2020\|EPI_ISL_475680\|2020-03- | ···················· | ···················· | ····················· | ···················· | ···················· | ···················· |
| D | 1496 | hCoV-19/USA/CA-CSMC67/2020\|EPI_ISL_475681\|2020-03- | ···················· | ···················· | ····················· | ···················· | ···················· | ···················· |
| D | 1497 | hCoV-19/USA/CA-CSMC68/2020\|EPI_ISL_475682\|2020-03- | ···················· | ·A·················· | ····················· | ···················· | ···················· | ···················· |
| D | 1498 | hCoV-19/USA/CA-CSMC69/2020\|EPI_ISL_475683\|2020-03- | ···················· | ···················· | ····················· | ···················· | ···················· | ···················· |
| D | 1499 | hCoV-19/USA/CA-CSMC7/2020\|EPI_ISL_475684\|2020-03-2 | ···················· | ···················· | ····················· | ···················· | ···················· | ···················· |
| D | 1500 | hCoV-19/USA/CA-CSMC70/2020\|EPI_ISL_475685\|2020-03- | ···················· | ···················· | ····················· | ···················· | ···················· | ···················· |
| D | 1501 | hCoV-19/USA/CA-CSMC71/2020\|EPI_ISL_475686\|2020-04- | ···················· | ···················· | ····················· | ···················· | ···················· | ···················· |
| D | 1502 | hCoV-19/USA/CA-CSMC72/2020\|EPI_ISL_475687\|2020-03- | ···················· | ···················· | ····················· | ···················· | ···················· | ···················· |
| D | 1503 | hCoV-19/USA/CA-CSMC73/2020\|EPI_ISL_475688\|2020-03- | ···················· | ···················· | ····················· | ···················· | ···················· | ···················· |
| D | 1504 | hCoV-19/USA/CA-CSMC74/2020\|EPI_ISL_475689\|2020-03- | ···················· | ···················· | ····················· | ···················· | ···················· | ···················· |
| D | 1505 | hCoV-19/USA/CA-CSMC75/2020\|EPI_ISL_475690\|2020-04- | ···················· | ···················· | ····················· | ···················· | ···················· | ···················· |
| D | 1506 | hCoV-19/USA/CA-CSMC76/2020\|EPI_ISL_475691\|2020-04- | ···················· | ···················· | ····················· | ···················· | ···················· | ···················· |
| D | 1507 | hCoV-19/USA/CA-CSMC77/2020\|EPI_ISL_475692\|2020-03- | ···················· | ···················· | ····················· | ···················· | ···················· | ···················· |
| D | 1508 | hCoV-19/USA/CA-CSMC79/2020\|EPI_ISL_475694\|2020-03- | ···················· | ···················· | ····················· | ···················· | ···················· | ···················· |
| D | 1509 | hCoV-19/USA/CA-CSMC8/2020\|EPI_ISL_475695\|2020-03-2 | ···················· | ···················· | ····················· | ···················· | ···················· | ···················· |
| D | 1510 | hCoV-19/USA/CA-CSMC81/2020\|EPI_ISL_475697\|2020-04- | ···················· | ···················· | ····················· | ···················· | ···················· | ···················· |
| D | 1511 | hCoV-19/USA/CA-CSMC82/2020\|EPI_ISL_475698\|2020-04- | ···················· | ···················· | ····················· | ···················· | ···················· | ···················· |
| D | 1512 | hCoV-19/USA/CA-CSMC83/2020\|EPI_ISL_475699\|2020-03- | ···················· | ···················· | ····················· | ···················· | ···················· | ···················· |
| D | 1513 | hCoV-19/Belgium/rega-0411441/2020\|EPI_ISL_476951\|2 | ···················· | ···················· | ····················· | ···················· | ···················· | ···················· |
| D | 1514 | hCoV-19/Belgium/rega-0411442/2020\|EPI_ISL_476952\|2 | ···················· | ···················· | ····················· | ···················· | ···················· | ···················· |
| D | 1515 | hCoV-19/Belgium/rega-0411443/2020\|EPI_ISL_476953\|2 | ···················· | ···················· | ····················· | ···················· | ···················· | ···················· |
| D | 1516 | hCoV-19/Belgium/rega-0411444/2020\|EPI_ISL_476954\|2 | ···················· | ···················· | ····················· | ···················· | ···················· | ···················· |
| D | 1517 | hCoV-19/Belgium/rega-0411445/2020\|EPI_ISL_476955\|2 | ···················· | ···················· | ····················· | ···················· | ···················· | ···················· |
| D | 1518 | hCoV-19/Belgium/rega-0411446/2020\|EPI_ISL_476956\|2 | ···················· | ···················· | ····················· | ···················· | ···················· | ···················· |
| D | 1519 | hCoV-19/Belgium/rega-0411447/2020\|EPI_ISL_476957\|2 | ···················· | ···················· | ····················· | ···················· | ···················· | ···················· |
| D | 1520 | hCoV-19/Belgium/rega-0411448/2020\|EPI_ISL_476958\|2 | ···················· | ···················· | ····················· | ···················· | ···················· | ···················· |
| D | 1521 | hCoV-19/Belgium/rega-0411449/2020\|EPI_ISL_476959\|2 | ···················· | ···················· | ····················· | ···················· | ···················· | ···················· |
| D | 1522 | hCoV-19/Belgium/rega-0411450/2020\|EPI_ISL_476960\|2 | ···················· | ···················· | ····················· | ···················· | ···················· | ···················· |
| D | 1523 | hCoV-19/Belgium/rega-0411451/2020\|EPI_ISL_476961\|2 | ···················· | ···················· | ····················· | ···················· | ···················· | ···················· |
| D | 1524 | hCoV-19/Belgium/rega-0411452/2020\|EPI_ISL_476962\|2 | ···················· | ···················· | ····················· | ···················· | ···················· | ···················· |
| D | 1525 | hCoV-19/Belgium/rega-0411453/2020\|EPI_ISL_476963\|2 | ···················· | ···················· | ····················· | ···················· | ···················· | ···················· |
| D | 1526 | hCoV-19/Belgium/rega-0411454/2020\|EPI_ISL_476964\|2 | ···················· | ···················· | ····················· | ···················· | ···················· | ···················· |
| D | 1527 | hCoV-19/Belgium/rega-0411455/2020\|EPI_ISL_476965\|2 | ···················· | ···················· | ····················· | ···················· | ···················· | ···················· |
| D | 1528 | hCoV-19/Belgium/rega-0411456/2020\|EPI_ISL_476966\|2 | ···················· | ···················· | ····················· | ···················· | ···················· | ···················· |
| D | 1529 | hCoV-19/Belgium/rega-0411457/2020\|EPI_ISL_476967\|2 | ···················· | ···················· | ····················· | ···················· | ···················· | ···················· |
| D | 1530 | hCoV-19/Belgium/rega-0411458/2020\|EPI_ISL_476968\|2 | ···················· | ···················· | ····················· | ···················· | ···················· | ···················· |
| D | 1531 | hCoV-19/Belgium/rega-0411459/2020\|EPI_ISL_476969\|2 | ···················· | ···················· | ····················· | ···················· | ···················· | ···················· |
| D | 1532 | hCoV-19/Belgium/rega-0429426/2020\|EPI_ISL_476995\|2 | ···················· | ···················· | ····················· | ···················· | ···················· | ···················· |
| D | 1533 | hCoV-19/Belgium/rega-0501480/2020\|EPI_ISL_476996\|2 | ···················· | ···················· | ····················· | ···················· | ···················· | ···················· |
| D | 1534 | hCoV-19/Belgium/rega-0502481/2020\|EPI_ISL_476997\|2 | ···················· | ···················· | ····················· | ···················· | ···················· | ···················· |
| D | 1535 | hCoV-19/Belgium/rega-0502482/2020\|EPI_ISL_476998\|2 | ···················· | ···················· | ····················· | ···················· | ···················· | ···················· |
| D | 1536 | hCoV-19/Belgium/rega-0503483/2020\|EPI_ISL_476999\|2 | ···················· | ···················· | ····················· | ···················· | ···················· | ···················· |
| D | 1537 | hCoV-19/Singapore/182/2020\|EPI_ISL_462300\|2020-03- | ···················· | ···················· | ····················· | ···················· | ···················· | ···················· |
| D | 1538 | hCoV-19/Singapore/183/2020\|EPI_ISL_462301\|2020-03- | ···················· | ···················· | ····················· | ···················· | ···················· | ···················· |
| D | 1539 | hCoV-19/Singapore/184/2020\|EPI_ISL_462302\|2020-04- | ···················· | ···················· | ····················· | ···················· | ···················· | ···················· |
| D | 1540 | hCoV-19/Singapore/185/2020\|EPI_ISL_462303\|2020-04- | ···················· | ···················· | ····················· | ···················· | ···················· | ···················· |
| D | 1541 | hCoV-19/Singapore/187/2020\|EPI_ISL_462305\|2020-02- | ···················· | ···················· | ····················· | ···················· | ···················· | ···················· |
| D | 1542 | hCoV-19/Singapore/188/2020\|EPI_ISL_462306\|2020-02- | ···················· | ···················· | ····················· | ···················· | ···················· | ···················· |
| D | 1543 | hCoV-19/Singapore/189/2020\|EPI_ISL_462307\|2020-04- | ···················· | ···················· | ····················· | ···················· | ···················· | ···················· |
| D | 1544 | hCoV-19/Singapore/190/2020\|EPI_ISL_462308\|2020-04- | ···················· | ···················· | ····················· | ···················· | ···················· | ···················· |
| D | 1545 | hCoV-19/Singapore/191/2020\|EPI_ISL_462309\|2020-04- | ···················· | ···················· | ····················· | ···················· | ···················· | ···················· |
| D | 1546 | hCoV-19/Singapore/236/2020\|EPI_ISL_462354\|2020-04- | ···················· | ···················· | ····················· | ···················· | ···················· | ···················· |
| D | 1547 | hCoV-19/Singapore/237/2020\|EPI_ISL_462355\|2020-04- | ···················· | ···················· | ····················· | ···················· | ···················· | ···················· |
| D | 1548 | hCoV-19/Singapore/238/2020\|EPI_ISL_462356\|2020-04- | ···················· | ···················· | ····················· | ···················· | ···················· | ···················· |
| D | 1549 | hCoV-19/Singapore/239/2020\|EPI_ISL_462357\|2020-04- | ···················· | ···················· | ····················· | ···················· | ···················· | ···················· |
| D | 1550 | hCoV-19/Singapore/240/2020\|EPI_ISL_462358\|2020-03- | ···················· | ···················· | ····················· | ···················· | ···················· | ···················· |
| D | 1551 | hCoV-19/Singapore/241/2020\|EPI_ISL_462359\|2020-03- | ···················· | ···················· | ····················· | ···················· | ···················· | ···················· |
| D | 1552 | hCoV-19/Singapore/242/2020\|EPI_ISL_462360\|2020-04- | ···················· | ···················· | ····················· | ···················· | ···················· | ···················· |
| D | 1553 | hCoV-19/Singapore/243/2020\|EPI_ISL_462361\|2020-04- | ···················· | ···················· | ····················· | ···················· | ···················· | ···················· |
| D | 1554 | hCoV-19/Singapore/251/2020\|EPI_ISL_462369\|2020-03- | ···················· | ···················· | ····················· | ···················· | ···················· | ···················· |
| D | 1555 | hCoV-19/Singapore/252/2020\|EPI_ISL_462370\|2020-03- | ···················· | ···················· | ····················· | ···················· | ···················· | ···················· |
| D | 1556 | hCoV-19/Singapore/253/2020\|EPI_ISL_462371\|2020-03- | ···················· | ···················· | ····················· | ···················· | ···················· | ···················· |
| D | 1557 | hCoV-19/Singapore/254/2020\|EPI_ISL_462372\|2020-05- | ···················· | ···················· | ····················· | ···················· | ···················· | ···················· |
| D | 1558 | hCoV-19/Singapore/255/2020\|EPI_ISL_462373\|2020-03- | ···················· | ···················· | ····················· | ···················· | ···················· | ···················· |
| D | 1559 | hCoV-19/Singapore/256/2020\|EPI_ISL_462374\|2020-04- | ···················· | ···················· | ····················· | ···················· | ···················· | ···················· |
| D | 1560 | hCoV-19/Singapore/257/2020\|EPI_ISL_462375\|2020-04- | ···················· | ···················· | ····················· | ···················· | ···················· | ···················· |
| D | 1561 | hCoV-19/USA/WA-S1120/2020\|EPI_ISL_463610\|2020-05-0 | ···················· | ···················· | ····················· | ···················· | ···················· | ···················· |
| D | 1562 | hCoV-19/USA/WA-S1121/2020\|EPI_ISL_463611\|2020-05-0 | ···················· | ···················· | ····················· | ···················· | ···················· | ···················· |
| D | 1563 | hCoV-19/USA/WA-S1122/2020\|EPI_ISL_463612\|2020-05-0 | ···················· | ···················· | ····················· | ···················· | ···················· | ···················· |
| D | 1564 | hCoV-19/USA/WA-S1123/2020\|EPI_ISL_463613\|2020-05-0 | ···················· | ···················· | ····················· | ···················· | ···················· | ···················· |
| D | 1565 | hCoV-19/USA/WA-S1124/2020\|EPI_ISL_463614\|2020-05-0 | ···················· | ···················· | ····················· | ···················· | ···················· | ···················· |
| D | 1566 | hCoV-19/USA/WA-S1125/2020\|EPI_ISL_463615\|2020-05-0 | ···················· | ···················· | ····················· | ···················· | ···················· | ···················· |
| D | 1567 | hCoV-19/USA/WA-S1128/2020\|EPI_ISL_463618\|2020-05-0 | ···················· | ···················· | ····················· | ···················· | ···················· | ···················· |
| D | 1568 | hCoV-19/USA/WA-S1129/2020\|EPI_ISL_463619\|2020-05-0 | ···················· | ···················· | ····················· | ···················· | ···················· | ···················· |
| D | 1569 | hCoV-19/USA/WA-S1130/2020\|EPI_ISL_463620\|2020-05-0 | ···················· | ···················· | ····················· | ···················· | ···················· | ···················· |
| D | 1570 | hCoV-19/USA/WA-S1131/2020\|EPI_ISL_463621\|2020-05-0 | ···················· | ···················· | ····················· | ···················· | ···················· | ···················· |
| D | 1571 | hCoV-19/USA/WA-S1132/2020\|EPI_ISL_463622\|2020-05-0 | ···················· | ···················· | ····················· | ···················· | ···················· | ···················· |
| D | 1572 | hCoV-19/USA/WA-S1133/2020\|EPI_ISL_463623\|2020-05-0 | ···················· | ···················· | ····················· | ···················· | ···················· | ···················· |

| | | | | | | | |
|---|---|---|---|---|---|---|---|
| D | 1573 hCoV-19/USA/WA-S1134/2020\|EPI_ISL_463624\|2020-05-0 | ···················· | ···················· | ···················· | ···················· | ···················· | ···················· |
| D | 1574 hCoV-19/USA/WA-S1135/2020\|EPI_ISL_463625\|2020-05-0 | ···················· | ···················· | ···················· | ···················· | ···················· | ···················· |
| D | 1575 hCoV-19/USA/WA-S1136/2020\|EPI_ISL_463626\|2020-05-0 | ···················· | ···················· | ···················· | ···················· | ···················· | ···················· |
| D | 1576 hCoV-19/USA/WA-S1137/2020\|EPI_ISL_463627\|2020-05-0 | ···················· | ···················· | ···················· | ···················· | ···················· | ···················· |
| D | 1577 hCoV-19/USA/WA-S1138/2020\|EPI_ISL_463628\|2020-05-0 | ···················· | ···················· | ···················· | ···················· | ···················· | ···················· |
| D | 1578 hCoV-19/USA/WA-S1139/2020\|EPI_ISL_463629\|2020-05-0 | ···················· | ···················· | ···················· | ···················· | ···················· | ···················· |
| D | 1579 hCoV-19/USA/WA-S1140/2020\|EPI_ISL_463630\|2020-05-0 | ···················· | ···················· | ···················· | ···················· | ···················· | ···················· |
| D | 1580 hCoV-19/USA/WA-S1142/2020\|EPI_ISL_463632\|2020-05-0 | ···················· | ···················· | ···················· | ···················· | ···················· | ···················· |
| D | 1581 hCoV-19/USA/WA-S1143/2020\|EPI_ISL_463633\|2020-05-0 | ···················· | ···················· | ···················· | ···················· | ···················· | ···················· |
| D | 1582 hCoV-19/USA/WA-S1144/2020\|EPI_ISL_463634\|2020-05-0 | ···················· | ···················· | ···················· | ···················· | ···················· | ···················· |
| D | 1583 hCoV-19/USA/WA-S1145/2020\|EPI_ISL_463635\|2020-05-0 | ···················· | ···················· | ···················· | ···················· | ···················· | ···················· |
| D | 1584 hCoV-19/USA/WA-S1146/2020\|EPI_ISL_463636\|2020-05-0 | ···················· | ···················· | ···················· | ···················· | ···················· | ···················· |
| D | 1585 hCoV-19/USA/WA-S1147/2020\|EPI_ISL_463637\|2020-05-0 | ···················· | ···················· | ···················· | ···················· | ···················· | ···················· |
| D | 1586 hCoV-19/USA/WA-S1148/2020\|EPI_ISL_463638\|2020-05-0 | ···················· | ···················· | ···················· | ···················· | ···················· | ···················· |
| D | 1587 hCoV-19/USA/WA-S1150/2020\|EPI_ISL_463640\|2020-05-0 | ···················· | ···················· | ···················· | ···················· | ···················· | ···················· |
| D | 1588 hCoV-19/USA/WA-S1196/2020\|EPI_ISL_463686\|2020-05-0 | ···················· | ···················· | ···················· | ···················· | ···················· | ···················· |
| D | 1589 hCoV-19/USA/WA-S1197/2020\|EPI_ISL_463687\|2020-05-0 | ···················· | ···················· | ···················· | ···················· | ···················· | ···················· |
| D | 1590 hCoV-19/USA/WA-S1198/2020\|EPI_ISL_463688\|2020-05-0 | ···················· | ·A·················· | ···················· | ···················· | ···················· | ···················· |
| D | 1591 hCoV-19/USA/WA-S1199/2020\|EPI_ISL_463689\|2020-05-0 | ···················· | ···················· | ···················· | ···················· | ···················· | ···················· |
| D | 1592 hCoV-19/USA/WA-S1200/2020\|EPI_ISL_463690\|2020-05-0 | ···················· | ···················· | ···················· | ···················· | ···················· | ···················· |
| D | 1593 hCoV-19/USA/WA-S1201/2020\|EPI_ISL_463691\|2020-05-0 | ···················· | ···················· | ···················· | ···················· | ···················· | ···················· |
| D | 1594 hCoV-19/USA/WA-S1202/2020\|EPI_ISL_463692\|2020-05-0 | ···················· | ···················· | ···················· | ···················· | ···················· | ···················· |
| D | 1595 hCoV-19/USA/WA-S1204/2020\|EPI_ISL_463694\|2020-05-0 | ···················· | ···················· | ···················· | ···················· | ···················· | ···················· |
| D | 1596 hCoV-19/USA/WA-S1206/2020\|EPI_ISL_463696\|2020-05-0 | ···················· | ···················· | ···················· | ···················· | ···················· | ···················· |
| D | 1597 hCoV-19/USA/WA-S1207/2020\|EPI_ISL_463697\|2020-05-0 | ···················· | ···················· | ···················· | ···················· | ···················· | ···················· |
| D | 1598 hCoV-19/USA/WA-S1208/2020\|EPI_ISL_463698\|2020-05-0 | ···················· | ···················· | ···················· | ···················· | ···················· | ···················· |
| D | 1599 hCoV-19/USA/WA-S1209/2020\|EPI_ISL_463699\|2020-05-0 | ···················· | ···················· | ···················· | ···················· | ···················· | ···················· |
| D | 1600 hCoV-19/England/201180343/2020\|EPI_ISL_464900\|2020 | ···················· | ···················· | ···················· | ···················· | ···················· | ···················· |
| D | 1601 hCoV-19/England/20118039804/2020\|EPI_ISL_464908\|20 | ···················· | ···················· | ···················· | ···················· | ···················· | ···················· |
| D | 1602 hCoV-19/England/20118040004/2020\|EPI_ISL_464909\|20 | ···················· | ···················· | ···················· | ···················· | ···················· | ···················· |
| D | 1603 hCoV-19/England/20118042504/2020\|EPI_ISL_464912\|20 | ···················· | ···················· | ···················· | ···················· | ···················· | ··NN················ |
| D | 1605 hCoV-19/England/20118050304/2020\|EPI_ISL_464920\|20 | ···················· | ···················· | ···················· | ···················· | ···················· | ···················· |
| D | 1606 hCoV-19/England/20118050404/2020\|EPI_ISL_464921\|20 | ···················· | ···················· | ···················· | ···················· | ···················· | ···················· |
| D | 1607 hCoV-19/England/20118050804/2020\|EPI_ISL_464923\|20 | ···················· | ···················· | ···················· | ···················· | ···················· | ···················· |
| D | 1608 hCoV-19/England/20118051204/2020\|EPI_ISL_464925\|20 | ···················· | ···················· | ···················· | ···················· | ···················· | ···················· |
| D | 1609 hCoV-19/England/20118054004/2020\|EPI_ISL_464926\|20 | ···················· | ···················· | ···················· | ···················· | ···················· | ···················· |
| D | 1610 hCoV-19/England/20118085604/2020\|EPI_ISL_464944\|20 | ···················· | ···················· | ···················· | ···················· | ···················· | ···················· |
| D | 1611 hCoV-19/England/20118087504/2020\|EPI_ISL_464946\|20 | ···················· | ···················· | ···················· | ···················· | ···················· | ···················· |
| D | 1612 hCoV-19/England/20118104104/2020\|EPI_ISL_464949\|20 | ···················· | ···················· | ···················· | ···················· | ···················· | ···················· |
| D | 1618 hCoV-19/England/20118112104/2020\|EPI_ISL_464957\|20 | ···················· | ···················· | ···················· | ···················· | ···················· | ···················· |
| D | 1619 hCoV-19/England/20118123004/2020\|EPI_ISL_464958\|20 | ···················· | ···················· | ···················· | ···················· | ···················· | ···················· |
| D | 1620 hCoV-19/England/20118123204/2020\|EPI_ISL_464959\|20 | ···················· | ···················· | ···················· | ···················· | ···················· | ···················· |
| D | 1623 hCoV-19/England/20118130804/2020\|EPI_ISL_464963\|20 | ···················· | ···················· | ···················· | ···················· | ···················· | ···················· |
| D | 1624 hCoV-19/England/20118142904/2020\|EPI_ISL_464971\|20 | ···················· | ···················· | ···················· | ···················· | ···················· | ···················· |
| D | 1625 hCoV-19/Spain/PV-IBV-001505/2020\|EPI_ISL_467170\|20 | ···················· | ···················· | ···················· | ···················· | ···················· | ···················· |
| D | 1626 hCoV-19/Spain/PV-IBV-001578/2020\|EPI_ISL_467171\|20 | ···················· | ···················· | ···················· | ···················· | ···················· | ···················· |
| D | 1627 hCoV-19/Spain/PV-IBV-001558/2020\|EPI_ISL_467172\|20 | ···················· | ···················· | ···················· | ···················· | ···················· | ···················· |
| D | 1628 hCoV-19/Spain/PV-IBV-001437/2020\|EPI_ISL_467173\|20 | ···················· | ···················· | ···················· | ···················· | ···················· | ···················· |
| D | 1629 hCoV-19/Spain/PV-IBV-001539/2020\|EPI_ISL_467174\|20 | ···················· | ···················· | ···················· | ···················· | ···················· | ···················· |
| D | 1630 hCoV-19/Spain/PV-IBV-001604/2020\|EPI_ISL_467175\|20 | ···················· | ···················· | ···················· | ···················· | ···················· | ···················· |
| D | 1631 hCoV-19/Spain/PV-IBV-001619/2020\|EPI_ISL_467176\|20 | ···················· | ···················· | ···················· | ···················· | ···················· | ···················· |
| D | 1632 hCoV-19/Spain/PV-IBV-001440/2020\|EPI_ISL_467177\|20 | ···················· | ···················· | ···················· | ···················· | ···················· | ···················· |
| D | 1633 hCoV-19/Spain/PV-IBV-001482/2020\|EPI_ISL_467178\|20 | ···················· | ···················· | ···················· | ···················· | ···················· | ···················· |
| D | 1634 hCoV-19/Spain/PV-IBV-001462/2020\|EPI_ISL_467179\|20 | ···················· | ···················· | ···················· | ···················· | ···················· | ···················· |
| D | 1635 hCoV-19/Spain/PV-IBV-001574/2020\|EPI_ISL_467180\|20 | ···················· | ···················· | ···················· | ···················· | ···················· | ···················· |
| D | 1636 hCoV-19/Spain/PV-IBV-001526/2020\|EPI_ISL_467181\|20 | ···················· | ···················· | ···················· | ···················· | ···················· | ···················· |
| D | 1637 hCoV-19/Spain/PV-IBV-001544/2020\|EPI_ISL_467182\|20 | ···················· | ···················· | ···················· | ···················· | ···················· | ···················· |
| D | 1639 hCoV-19/Spain/MD-IBV-003705/2020\|EPI_ISL_467184\|20 | ···················· | ···················· | ···················· | ···················· | ···················· | ···················· |
| D | 1641 hCoV-19/Spain/MD-IBV-003687/2020\|EPI_ISL_467186\|20 | ···················· | ···················· | ···················· | ···················· | ···················· | ···················· |
| D | 1642 hCoV-19/Spain/MD-IBV-003669/2020\|EPI_ISL_467188\|20 | ···················· | ···················· | ···················· | ···················· | ···················· | ···················· |
| D | 1643 hCoV-19/Spain/MD-IBV-003663/2020\|EPI_ISL_467189\|20 | ···················· | ···················· | ···················· | ···················· | ···················· | ···················· |
| D | 1644 hCoV-19/Spain/MD-IBV-003722/2020\|EPI_ISL_467190\|20 | ···················· | ···················· | ···················· | ···················· | ···················· | ···················· |
| D | 1647 hCoV-19/Spain/MD-IBV-003667/2020\|EPI_ISL_467193\|20 | ···················· | ···················· | ···················· | ···················· | ···················· | ···················· |
| D | 1648 hCoV-19/Spain/MD-IBV-003719/2020\|EPI_ISL_467194\|20 | ···················· | ···················· | ···················· | ···················· | ···················· | ···················· |
| D | 1649 hCoV-19/Spain/MD-IBV-003724/2020\|EPI_ISL_467195\|20 | ···················· | ···················· | ···················· | ···················· | ···················· | ···················· |
| D | 1652 hCoV-19/Spain/MD-IBV-003685/2020\|EPI_ISL_467198\|20 | ···················· | ···················· | ···················· | ···················· | ···················· | ···················· |
| D | 1653 hCoV-19/Spain/MD-IBV-003708/2020\|EPI_ISL_467199\|20 | ···················· | ···················· | ···················· | ···················· | ···················· | ···················· |
| D | 1654 hCoV-19/Wales/PHWC-2D52C/2020\|EPI_ISL_474316\|2020- | ···················· | ···················· | ···················· | ···················· | ···················· | ···················· |
| D | 1655 hCoV-19/Wales/PHWC-2DEC7/2020\|EPI_ISL_474318\|2020- | ···················· | ···················· | ···················· | ···················· | ···················· | ···················· |
| D | 1656 hCoV-19/Wales/PHWC-30FBF/2020\|EPI_ISL_474322\|2020- | ···················· | ···················· | ···················· | ···················· | ···················· | ···················· |
| D | 1657 hCoV-19/Wales/PHWC-3126A/2020\|EPI_ISL_474323\|2020- | ···················· | ···················· | ···················· | ···················· | ···················· | ···················· |
| D | 1658 hCoV-19/USA/CA-CSMC123/2020\|EPI_ISL_475600\|2020-03 | ···················· | ···················· | ···················· | ···················· | ···················· | ···················· |
| D | 1659 hCoV-19/USA/CA-CSMC124/2020\|EPI_ISL_475601\|2020-04 | ···················· | ···················· | ···················· | ···················· | ···················· | ···················· |
| D | 1660 hCoV-19/USA/CA-CSMC125/2020\|EPI_ISL_475602\|2020-04 | ···················· | ···················· | ···················· | ···················· | ···················· | ···················· |
| D | 1661 hCoV-19/USA/CA-CSMC126/2020\|EPI_ISL_475603\|2020-04 | ···················· | ···················· | ···················· | ···················· | ···················· | ···················· |
| D | 1662 hCoV-19/USA/CA-CSMC127/2020\|EPI_ISL_475604\|2020-04 | ···················· | ···················· | ···················· | ···················· | ···················· | ···················· |
| D | 1663 hCoV-19/USA/CA-CSMC128/2020\|EPI_ISL_475605\|2020-04 | ···················· | ···················· | ···················· | ···················· | ···················· | ···················· |
| D | 1664 hCoV-19/USA/CA-CSMC129/2020\|EPI_ISL_475606\|2020-04 | ···················· | ···················· | ···················· | ···················· | ···················· | ···················· |

D 1665 hCoV-19/USA/CA-CSMC13/2020|EPI_ISL_475607|2020-03- ···················· ···················· ····················· ···················· ···················· ····················
D 1666 hCoV-19/USA/CA-CSMC130/2020|EPI_ISL_475608|2020-03 ···················· ···················· ····················· ···················· ···················· ····················
D 1667 hCoV-19/USA/CA-CSMC131/2020|EPI_ISL_475609|2020-03 ···················· ···················· ····················· ···················· ···················· ····················
D 1668 hCoV-19/USA/CA-CSMC132/2020|EPI_ISL_475610|2020-03 ···················· ···················· ····················· ···················· ···················· ····················
D 1669 hCoV-19/USA/CA-CSMC133/2020|EPI_ISL_475611|2020-03 ···················· ···················· ····················· ···················· ···················· ····················
D 1670 hCoV-19/USA/CA-CSMC134/2020|EPI_ISL_475612|2020-03 ···················· ···················· ····················· ···················· ···················· ····················
D 1671 hCoV-19/USA/CA-CSMC135/2020|EPI_ISL_475613|2020-03 ···················· ···················· ····················· ···················· ···················· ····················
D 1672 hCoV-19/USA/CA-CSMC136/2020|EPI_ISL_475614|2020-04 ···················· ···················· ····················· ···················· ···················· ····················
D 1673 hCoV-19/USA/CA-CSMC137/2020|EPI_ISL_475615|2020-04 ···················· ···················· ····················· ···················· ···················· ····················
D 1674 hCoV-19/USA/CA-CSMC138/2020|EPI_ISL_475616|2020-04 ···················· ···················· ····················· ···················· ···················· ····················
D 1675 hCoV-19/USA/CA-CSMC139/2020|EPI_ISL_475617|2020-04 ···················· ···················· ····················· ···················· ···················· ····················
D 1676 hCoV-19/USA/CA-CSMC14/2020|EPI_ISL_475618|2020-03- ···················· ···················· ····················· ···················· ···················· ····················
D 1677 hCoV-19/USA/CA-CSMC140/2020|EPI_ISL_475619|2020-04 ···················· ···················· ····················· ···················· ···················· ····················
D 1678 hCoV-19/USA/CA-CSMC3/2020|EPI_ISL_475640|2020-03-2 ···················· ···················· ····················· ···················· ···················· ····················
D 1679 hCoV-19/USA/CA-CSMC30/2020|EPI_ISL_475641|2020-03- ···················· ···················· ····················· ···················· ···················· ····················
D 1680 hCoV-19/USA/CA-CSMC32/2020|EPI_ISL_475643|2020-04- ···················· ···················· ····················· ···················· ···················· ····················
D 1681 hCoV-19/USA/CA-CSMC33/2020|EPI_ISL_475644|2020-04- ···················· ···················· ····················· ···················· ···················· ····················
D 1682 hCoV-19/USA/CA-CSMC34/2020|EPI_ISL_475645|2020-04- ···················· ···················· ····················· ···················· ···················· ····················
D 1683 hCoV-19/USA/CA-CSMC35/2020|EPI_ISL_475646|2020-04- ···················· ···················· ····················· ···················· ···················· ····················
D 1684 hCoV-19/USA/CA-CSMC36/2020|EPI_ISL_475647|2020-04- ···················· ···················· ····················· ···················· ···················· ····················
D 1685 hCoV-19/USA/CA-CSMC37/2020|EPI_ISL_475648|2020-04- ···················· ···················· ····················· ···················· ···················· ····················
D 1686 hCoV-19/USA/CA-CSMC39/2020|EPI_ISL_475650|2020-04- ···················· ···················· ····················· ···················· ···················· ····················
D 1687 hCoV-19/USA/CA-CSMC4/2020|EPI_ISL_475651|2020-03-2 ···················· ···················· ····················· ···················· ···················· ····················
D 1688 hCoV-19/USA/CA-CSMC40/2020|EPI_ISL_475652|2020-04- ···················· ···················· ····················· ···················· ···················· ····················
D 1689 hCoV-19/USA/CA-CSMC41/2020|EPI_ISL_475653|2020-03- ···················· ···················· ····················· ···················· ···················· ····················
D 1690 hCoV-19/USA/CA-CSMC42/2020|EPI_ISL_475654|2020-04- ···················· ···················· ····················· ···················· ···················· ····················
D 1691 hCoV-19/USA/CA-CSMC43/2020|EPI_ISL_475655|2020-04- ···················· ···················· ····················· ···················· ···················· ····················
D 1692 hCoV-19/USA/CA-CSMC44/2020|EPI_ISL_475656|2020-03- ···················· ···················· ····················· ···················· ···················· ····················
D 1693 hCoV-19/USA/CA-CSMC45/2020|EPI_ISL_475657|2020-03- ···················· ···················· ····················· ···················· ···················· ····················
D 1694 hCoV-19/USA/CA-CSMC46/2020|EPI_ISL_475658|2020-04- ···················· ···················· ····················· ···················· ···················· ····················
D 1695 hCoV-19/USA/CA-CSMC47/2020|EPI_ISL_475659|2020-04- ···················· ···················· ····················· ···················· ···················· ····················
D 1696 hCoV-19/USA/CA-CSMC48/2020|EPI_ISL_475660|2020-04- ···················· ···················· ····················· ···················· ···················· ····················
D 1697 hCoV-19/USA/CA-CSMC49/2020|EPI_ISL_475661|2020-04- ···················· ···················· ····················· ···················· ···················· ····················
D 1698 hCoV-19/USA/CA-CSMC5/2020|EPI_ISL_475662|2020-04-1 ···················· ···················· ····················· ···················· ···················· ····················
D 1699 hCoV-19/USA/CA-CSMC50/2020|EPI_ISL_475663|2020-04- ···················· ···················· ····················· ···················· ···················· ····················
D 1700 hCoV-19/USA/UN-UW-629/2020|EPI_ISL_476910|2020-03- ···················· ···················· ····················· ···················· ···················· ····················
D 1701 hCoV-19/USA/WA-UW-631/2020|EPI_ISL_476911|2020-03- ···················· ···················· ···········T········· ···················· ···················· ····················
D 1702 hCoV-19/USA/CT-UW-645/2020|EPI_ISL_476912|2020-03- ···················· ···················· ····················· ···················· ···················· ····················
D 1703 hCoV-19/USA/CT-UW-649/2020|EPI_ISL_476914|2020-03- ···················· ···················· ····················· ···················· ···················· ····················
D 1704 hCoV-19/USA/CT-UW-650/2020|EPI_ISL_476915|2020-03- ···················· ···················· ····················· ···················· ···················· ····················
D 1705 hCoV-19/USA/CT-UW-651/2020|EPI_ISL_476916|2020-03- ···················· ···················· ····················· ···················· ···················· ····················
D 1706 hCoV-19/USA/CT-UW-652/2020|EPI_ISL_476917|2020-03- ···················· ···················· ····················· ···················· ···················· ····················
D 1707 hCoV-19/USA/CT-UW-653/2020|EPI_ISL_476918|2020-03- ···················· ···················· ····················· ···················· ···················· ····················
D 1708 hCoV-19/USA/UN-UW-659/2020|EPI_ISL_476919|2020-03- ···················· ···················· ····················· ···················· ···················· ····················
D 1709 hCoV-19/USA/WA-UW-661/2020|EPI_ISL_476920|2020-03- ···················· ···················· ····················· ···················· ···················· ····················
D 1710 hCoV-19/USA/WA-UW-662/2020|EPI_ISL_476921|2020-03- ···················· ···················· ····················· ···················· ···················· ····················
D 1711 hCoV-19/USA/UN-UW-666/2020|EPI_ISL_476922|2020-03- ···················· ···················· ····················· ···················· ···················· ····················
D 1712 hCoV-19/USA/UN-UW-667/2020|EPI_ISL_476923|2020-03- ···················· ···················· ····················· ···················· ···················· ····················
D 1713 hCoV-19/USA/WA-UW-669/2020|EPI_ISL_476924|2020-03- ···················· ···················· ····················· ···················· ···················· ····················
D 1714 hCoV-19/USA/WA-UW-671/2020|EPI_ISL_476925|2020-03- ···················· ···················· ····················· ···················· ···················· ····················
D 1715 hCoV-19/USA/UN-UW-672/2020|EPI_ISL_476926|2020-03- ···················· ···················· ····················· ···················· ···················· ····················
D 1716 hCoV-19/USA/WA-UW-674/2020|EPI_ISL_476927|2020-03- ···················· ···················· ····················· ···················· ···················· ····················
D 1717 hCoV-19/USA/WA-UW-676/2020|EPI_ISL_476928|2020-03- ···················· ···················· ····················· ···················· ···················· ····················
D 1718 hCoV-19/USA/WA-UW-681/2020|EPI_ISL_476929|2020-03- ···················· ···················· ····················· ···················· ···················· ····················
D 1719 hCoV-19/USA/WA-UW-683/2020|EPI_ISL_476930|2020-03- ···················· ···················· ····················· ···················· ···················· ····················
D 1720 hCoV-19/USA/CT-UW-685/2020|EPI_ISL_476931|2020-03- ···················· ···················· ····················· ···················· ···················· ····················
D 1721 hCoV-19/USA/UN-UW-686/2020|EPI_ISL_476932|2020-03- ···················· ···················· ····················· ···················· ···················· ····················
D 1722 hCoV-19/USA/WA-UW-687/2020|EPI_ISL_476933|2020-03- ···················· ···················· ····················· ···················· ···················· ····················
D 1723 hCoV-19/USA/WA-UW-696/2020|EPI_ISL_476935|2020-03- ···················· ···················· ····················· ···················· ···················· ····················
D 1724 hCoV-19/USA/WA-UW-753/2020|EPI_ISL_476936|2020-03- ···················· ···················· ····················· ···················· ···················· ····················
D 1725 hCoV-19/USA/WA-UW-862/2020|EPI_ISL_476937|2020-03- ···················· ···················· ····················· ···················· ···················· ····················
D 1726 hCoV-19/USA/WA-UW-864/2020|EPI_ISL_476938|2020-03- ···················· ···················· ····················· ···················· ···················· ····················
D 1727 hCoV-19/USA/WA-UW-865/2020|EPI_ISL_476939|2020-03- ···················· ···················· ····················· ···················· ···················· ····················
D 1728 hCoV-19/USA/WA-UW-896/2020|EPI_ISL_476940|2020-03- ···················· ···················· ····················· ···················· ···················· ····················
D 1729 hCoV-19/Belgium/rega-0409431/2020|EPI_ISL_476941|2 ···················· ···················· ····················· ···················· ···················· ····················
D 1730 hCoV-19/Belgium/rega-0409433/2020|EPI_ISL_476942|2 ···················· ···················· ····················· ···················· ···················· ····················
D 1731 hCoV-19/Belgium/rega-0409434/2020|EPI_ISL_476943|2 ···················· ···················· ····················· ···················· ···················· ····················
D 1732 hCoV-19/Belgium/rega-0409435/2020|EPI_ISL_476944|2 ···················· ···················· ····················· ···················· ···················· ····················
D 1733 hCoV-19/Belgium/rega-0409436/2020|EPI_ISL_476945|2 ···················· ···················· ····················· ···················· ···················· ····················
D 1734 hCoV-19/Belgium/rega-0409437/2020|EPI_ISL_476946|2 ···················· ···················· ····················· ···················· ···················· ····················
D 1735 hCoV-19/Belgium/rega-0409438/2020|EPI_ISL_476947|2 ···················· ···················· ····················· ···················· ···················· ····················
D 1736 hCoV-19/Belgium/rega-0409439/2020|EPI_ISL_476948|2 ···················· ···················· ····················· ···················· ···················· ····················
D 1737 hCoV-19/Belgium/rega-0409497/2020|EPI_ISL_476949|2 ···················· ···················· ····················· ···················· ···················· ····················
D 1738 hCoV-19/Belgium/rega-0411440/2020|EPI_ISL_476950|2 ···················· ···················· ····················· ···················· ···················· ····················
D 1739 hCoV-19/Belgium/rega-0411460/2020|EPI_ISL_476970|2 ···················· ···················· ····················· ···················· ···················· ····················
D 1740 hCoV-19/Belgium/rega-0411461/2020|EPI_ISL_476971|2 ···················· ···················· ····················· ···················· ···················· ····················
D 1741 hCoV-19/Belgium/rega-0411462/2020|EPI_ISL_476972|2 ···················· ···················· ····················· ···················· ···················· ····················
D 1742 hCoV-19/Belgium/rega-0411463/2020|EPI_ISL_476973|2 ···················· ···················· ····················· ···················· ···················· ····················

| | | | | | | | | |
|---|---|---|---|---|---|---|---|---|
| D | 1743 | hCoV-19/Belgium/rega-0411464/2020\|EPI_ISL_476974\|2 | .................... | .................... | .................... | .................... | .................... | .................... |
| D | 1744 | hCoV-19/Belgium/rega-0411465/2020\|EPI_ISL_476975\|2 | .................... | .................... | .................... | .................... | .................... | .................... |
| D | 1745 | hCoV-19/Belgium/rega-0411466/2020\|EPI_ISL_476976\|2 | .................... | .................... | .................... | .................... | .................... | .................... |
| D | 1746 | hCoV-19/Belgium/rega-0411467/2020\|EPI_ISL_476977\|2 | .................... | .................... | .................... | .................... | .................... | .................... |
| D | 1747 | hCoV-19/Belgium/rega-0411468/2020\|EPI_ISL_476978\|2 | .................... | .................... | .................... | .................... | .................... | .................... |
| D | 1748 | hCoV-19/Belgium/rega-0411469/2020\|EPI_ISL_476979\|2 | .................... | .................... | .................... | .................... | .................... | .................... |
| D | 1749 | hCoV-19/Belgium/rega-0411470/2020\|EPI_ISL_476980\|2 | .................... | .................... | .................... | .................... | .................... | .................... |
| D | 1750 | hCoV-19/Belgium/rega-0411471/2020\|EPI_ISL_476981\|2 | .................... | .................... | .................... | .................... | .................... | .................... |
| D | 1751 | hCoV-19/Belgium/rega-0411472/2020\|EPI_ISL_476982\|2 | .................... | .................... | .................... | .................... | .................... | .................... |
| D | 1752 | hCoV-19/Belgium/rega-0414473/2020\|EPI_ISL_476983\|2 | .................... | .................... | .................... | .................... | .................... | .................... |
| D | 1753 | hCoV-19/Belgium/rega-0414474/2020\|EPI_ISL_476984\|2 | .................... | .................... | .................... | .................... | .................... | .................... |
| D | 1754 | hCoV-19/Belgium/rega-0414475/2020\|EPI_ISL_476985\|2 | .................... | .................... | .................... | .................... | .................... | .................... |
| D | 1755 | hCoV-19/Belgium/rega-0414476/2020\|EPI_ISL_476986\|2 | .................... | .................... | .................... | .................... | .................... | .................... |
| D | 1756 | hCoV-19/Belgium/rega-0414477/2020\|EPI_ISL_476987\|2 | .................... | .................... | .................... | .................... | .................... | .................... |
| D | 1757 | hCoV-19/Belgium/rega-0414478/2020\|EPI_ISL_476988\|2 | .................... | .................... | .................... | .................... | .................... | .................... |
| D | 1758 | hCoV-19/Belgium/rega-0414479/2020\|EPI_ISL_476989\|2 | .................... | .................... | .................... | .................... | .................... | .................... |
| D | 1759 | hCoV-19/Belgium/rega-0418422/2020\|EPI_ISL_476990\|2 | .................... | .................... | .................... | .................... | .................... | .................... |
| D | 1760 | hCoV-19/Belgium/rega-0421423/2020\|EPI_ISL_476991\|2 | .................... | .................... | .................... | .................... | .................... | .................... |
| D | 1761 | hCoV-19/Belgium/rega-0427424/2020\|EPI_ISL_476992\|2 | .................... | .................... | .................... | .................... | .................... | .................... |
| D | 1762 | hCoV-19/Belgium/rega-0428425/2020\|EPI_ISL_476993\|2 | .................... | .................... | .................... | .................... | .................... | .................... |
| D | 1763 | hCoV-19/Belgium/rega-0428427/2020\|EPI_ISL_476994\|2 | .................... | .................... | .................... | .................... | .................... | .................... |
| D | 1764 | hCoV-19/England/OXON-B24E3/2020\|EPI_ISL_479150\|202 | .................... | .................... | .................... | .................... | .................... | .................... |
| D | 1765 | hCoV-19/England/OXON-B250E/2020\|EPI_ISL_479152\|202 | .................... | .................... | .................... | .................... | .................... | .................... |
| D | 1766 | hCoV-19/England/OXON-B253B/2020\|EPI_ISL_479153\|202 | .................... | .................... | .................... | .................... | .................... | .................... |
| D | 1767 | hCoV-19/England/OXON-B254A/2020\|EPI_ISL_479154\|202 | .................... | .................... | .................... | .................... | .................... | .................... |
| D | 1768 | hCoV-19/England/OXON-B2559/2020\|EPI_ISL_479155\|202 | .................... | .................... | .................... | .................... | .................... | .................... |
| D | 1769 | hCoV-19/England/OXON-B2586/2020\|EPI_ISL_479156\|202 | .................... | .................... | .................... | .................... | .................... | .................... |
| D | 1770 | hCoV-19/England/OXON-B25B3/2020\|EPI_ISL_479157\|202 | .................... | .................... | .................... | .................... | .................... | .................... |
| D | 1771 | hCoV-19/England/OXON-B25C2/2020\|EPI_ISL_479158\|202 | .................... | .................... | .................... | .................... | .................... | .................... |
| D | 1772 | hCoV-19/England/OXON-B25FF/2020\|EPI_ISL_479160\|202 | .................... | .................... | .................... | .................... | .................... | .................... |
| D | 1773 | hCoV-19/England/OXON-B2647/2020\|EPI_ISL_479162\|202 | .................... | .................... | .................... | .................... | .................... | .................... |
| D | 1774 | hCoV-19/England/OXON-B2692/2020\|EPI_ISL_479165\|202 | .................... | .................... | .................... | .................... | .................... | .................... |
| D | 1775 | hCoV-19/England/OXON-B26B0/2020\|EPI_ISL_479166\|202 | .................... | .................... | .................... | .................... | .................... | .................... |
| D | 1776 | hCoV-19/England/OXON-B26ED/2020\|EPI_ISL_479167\|202 | .................... | .A.................. | .................... | .................... | .................... | .................... |
| D | 1777 | hCoV-19/England/OXON-B26FC/2020\|EPI_ISL_479168\|202 | .................... | .................... | .................... | .................... | .................... | .................... |
| D | 1778 | hCoV-19/England/OXON-B2708/2020\|EPI_ISL_479169\|202 | .................... | .................... | .................... | .................... | .................... | .................... |
| D | 1779 | hCoV-19/England/OXON-B2717/2020\|EPI_ISL_479170\|202 | .................... | .................... | .................... | .................... | .................... | .................... |
| D | 1780 | hCoV-19/England/OXON-B2726/2020\|EPI_ISL_479171\|202 | .................... | .............A...... | .................... | .................... | .................... | .................... |
| D | 1781 | hCoV-19/England/OXON-B2735/2020\|EPI_ISL_479172\|202 | .................... | .................... | .................... | .................... | .................... | .................... |
| D | 1782 | hCoV-19/England/OXON-B2744/2020\|EPI_ISL_479173\|202 | .................... | .................... | .................... | .................... | .................... | .................... |
| D | 1783 | hCoV-19/England/PORT-2D12C4/2020\|EPI_ISL_479176\|20 | .................... | .................... | .................... | .................... | .................... | .................... |
| D | 1784 | hCoV-19/England/PORT-2D12D3/2020\|EPI_ISL_479177\|20 | .................... | .................... | .................... | .................... | .................... | .................... |
| D | 1785 | hCoV-19/England/PORT-2D12E2/2020\|EPI_ISL_479178\|20 | .................... | .................... | .................... | .................... | .................... | .................... |
| D | 1786 | hCoV-19/England/PORT-2D12F1/2020\|EPI_ISL_479179\|20 | .................... | .................... | .................... | .................... | .................... | .................... |
| D | 1787 | hCoV-19/England/PORT-2D13A3/2020\|EPI_ISL_479190\|20 | .................... | .................... | .................... | .................... | .................... | .................... |
| D | 1788 | hCoV-19/England/PORT-2D13B2/2020\|EPI_ISL_479191\|20 | .................... | .................... | .................... | .................... | .................... | .................... |
| D | 1789 | hCoV-19/England/PORT-2D13D0/2020\|EPI_ISL_479192\|20 | .................... | .................... | .................... | .................... | .................... | .................... |
| D | 1790 | hCoV-19/England/PORT-2D13EF/2020\|EPI_ISL_479193\|20 | .................... | .................... | .................... | .................... | .................... | .................... |
| D | 1791 | hCoV-19/England/PORT-2D13FE/2020\|EPI_ISL_479194\|20 | .................... | .................... | .................... | .................... | .................... | .................... |
| D | 1792 | hCoV-19/England/SHEF-C1830/2020\|EPI_ISL_479198\|202 | .................... | .................... | .................... | .................... | .................... | .................... |
| D | 1793 | hCoV-19/USA/WA-S1110/2020\|EPI_ISL_463600\|2020-05-0 | .................... | .................... | .................... | .................... | .................... | .................... |
| D | 1794 | hCoV-19/USA/WA-S1111/2020\|EPI_ISL_463601\|2020-05-0 | .................... | .................... | .................... | .................... | .................... | .................... |
| D | 1795 | hCoV-19/USA/WA-S1112/2020\|EPI_ISL_463602\|2020-05-0 | .................... | .................... | .................... | .................... | .................... | .................... |
| D | 1796 | hCoV-19/USA/WA-S1113/2020\|EPI_ISL_463603\|2020-05-0 | .................... | .................... | .................... | .................... | .................... | .................... |
| D | 1797 | hCoV-19/USA/WA-S1114/2020\|EPI_ISL_463604\|2020-05-0 | .................... | .................... | .................... | .................... | .................... | .................... |
| D | 1798 | hCoV-19/USA/WA-S1115/2020\|EPI_ISL_463605\|2020-05-0 | .................... | .................... | .................... | .................... | .................... | .................... |
| D | 1799 | hCoV-19/USA/WA-S1117/2020\|EPI_ISL_463607\|2020-05-0 | .................... | .................... | .................... | .................... | .................... | .................... |
| D | 1800 | hCoV-19/USA/WA-S1118/2020\|EPI_ISL_463608\|2020-05-0 | .................... | .................... | .................... | .................... | .................... | .................... |
| D | 1801 | hCoV-19/USA/WA-S1119/2020\|EPI_ISL_463609\|2020-05-0 | .................... | .................... | .................... | .................... | .................... | .................... |
| D | 1802 | hCoV-19/USA/WA-S1126/2020\|EPI_ISL_463616\|2020-05-0 | .................... | .................... | .................... | .................... | .................... | .................... |
| D | 1803 | hCoV-19/USA/WA-S1127/2020\|EPI_ISL_463617\|2020-05-0 | .................... | .................... | .................... | .................... | .................... | .................... |
| D | 1804 | hCoV-19/England/20118059904/2020\|EPI_ISL_464932\|20 | .................... | .................... | .................... | .................... | .................... | .................... |
| D | 1805 | hCoV-19/England/20118061804/2020\|EPI_ISL_464936\|20 | .................... | .................... | .................... | .................... | .................... | .................... |
| D | 1807 | hCoV-19/England/20118070604/2020\|EPI_ISL_464939\|20 | .................... | .................... | .................... | .................... | .................... | .................... |
| D | 1809 | hCoV-19/England/20118104004/2020\|EPI_ISL_464948\|20 | .................... | .................... | .................... | .................... | .................... | .................... |
| D | 1810 | hCoV-19/Spain/PV-IBV-001520/2020\|EPI_ISL_467100\|20 | .................... | .................... | .................... | .................... | .................... | .................... |
| D | 1811 | hCoV-19/Spain/PV-IBV-001467/2020\|EPI_ISL_467101\|20 | .................... | .................... | .................... | .................... | .................... | .................... |
| D | 1812 | hCoV-19/Spain/PV-IBV-001587/2020\|EPI_ISL_467103\|20 | .................... | .................... | .................... | .................... | .................... | .................... |
| D | 1813 | hCoV-19/Spain/PV-IBV-001583/2020\|EPI_ISL_467104\|20 | .................... | .................... | .................... | .................... | .................... | .................... |
| D | 1814 | hCoV-19/Spain/PV-IBV-001575/2020\|EPI_ISL_467105\|20 | .................... | .................... | .................... | .................... | .................... | .................... |
| D | 1815 | hCoV-19/Spain/PV-IBV-001522/2020\|EPI_ISL_467106\|20 | .................... | .................... | .................... | .................... | .................... | .................... |
| D | 1816 | hCoV-19/Spain/PV-IBV-001559/2020\|EPI_ISL_467107\|20 | .................... | .................... | .................... | .................... | .................... | .................... |
| D | 1817 | hCoV-19/Spain/PV-IBV-001438/2020\|EPI_ISL_467108\|20 | .................... | .................... | .................... | .................... | ..N................. | .................... |
| D | 1819 | hCoV-19/Spain/PV-IBV-001547/2020\|EPI_ISL_467110\|20 | .................... | .................... | .................... | .................... | .................... | .................... |
| D | 1820 | hCoV-19/Spain/PV-IBV-001556/2020\|EPI_ISL_467111\|20 | .................... | .................... | .................... | .................... | .................... | .................... |
| D | 1821 | hCoV-19/Spain/PV-IBV-001566/2020\|EPI_ISL_467113\|20 | .................... | .................... | .................... | .................... | .................... | .................... |
| D | 1822 | hCoV-19/Spain/PV-IBV-001553/2020\|EPI_ISL_467114\|20 | .................... | .................... | .................... | .................... | .................... | .................... |
| D | 1823 | hCoV-19/Spain/PV-IBV-001443/2020\|EPI_ISL_467115\|20 | .................... | .................... | .................... | .................... | .................... | .................... |

| | | | | | | | | | |
|---|---|---|---|---|---|---|---|---|---|
| D | 1824 | hCoV-19/Spain/PV-IBV-001560/2020\|EPI_ISL_467116\|20 | ···················· | ···················· | ···················· | ···················· | ···················· | ···················· | |
| D | 1825 | hCoV-19/Spain/PV-IBV-001468/2020\|EPI_ISL_467117\|20 | ···················· | ···················· | ···················· | ···················· | ···················· | ···················· | |
| D | 1826 | hCoV-19/Spain/PV-IBV-001473/2020\|EPI_ISL_467118\|20 | ···················· | ···················· | ···················· | ···················· | ···················· | ···················· | |
| D | 1827 | hCoV-19/Spain/PV-IBV-001540/2020\|EPI_ISL_467120\|20 | ···················· | ···················· | ···················· | ···················· | ···················· | ···················· | |
| D | 1829 | hCoV-19/Spain/PV-IBV-001512/2020\|EPI_ISL_467122\|20 | ···················· | ···················· | ···················· | ···················· | ···················· | ···················· | |
| D | 1830 | hCoV-19/Spain/PV-IBV-001523/2020\|EPI_ISL_467123\|20 | ···················· | ···················· | ···················· | ···················· | ···················· | ···················· | |
| D | 1831 | hCoV-19/Spain/PV-IBV-001580/2020\|EPI_ISL_467124\|20 | ···················· | ·A·················· | ···················· | ···················· | ···················· | ···················· | |
| D | 1832 | hCoV-19/Spain/PV-IBV-001530/2020\|EPI_ISL_467125\|20 | ···················· | ···················· | ···················· | ···················· | ···················· | ···················· | |
| D | 1833 | hCoV-19/Spain/PV-IBV-001601/2020\|EPI_ISL_467126\|20 | ···················· | ···················· | ···················· | ···················· | ···················· | ···················· | |
| D | 1834 | hCoV-19/Spain/PV-IBV-001465/2020\|EPI_ISL_467127\|20 | ···················· | ···················· | ···················· | ···················· | ···················· | ···················· | |
| D | 1835 | hCoV-19/Spain/PV-IBV-001594/2020\|EPI_ISL_467142\|20 | ···················· | ···················· | ···················· | ···················· | ···················· | ···················· | |
| D | 1836 | hCoV-19/Spain/PV-IBV-001435/2020\|EPI_ISL_467143\|20 | ···················· | ···················· | ···················· | ···················· | ···················· | ···················· | |
| D | 1837 | hCoV-19/Spain/PV-IBV-001535/2020\|EPI_ISL_467144\|20 | ···················· | ···················· | ···················· | ···················· | ···················· | ···················· | |
| D | 1838 | hCoV-19/Spain/PV-IBV-001455/2020\|EPI_ISL_467145\|20 | ···················· | ···················· | ···················· | ···················· | ···················· | ···················· | |
| D | 1839 | hCoV-19/Spain/PV-IBV-001459/2020\|EPI_ISL_467146\|20 | ···················· | ···················· | ···················· | ···················· | ···················· | ···················· | |
| D | 1840 | hCoV-19/Spain/PV-IBV-001576/2020\|EPI_ISL_467147\|20 | ···················· | ···················· | ···················· | ···················· | ···················· | ···················· | |
| D | 1841 | hCoV-19/Spain/PV-IBV-001445/2020\|EPI_ISL_467148\|20 | ···················· | ·A·················· | ···················· | ···················· | ···················· | ···················· | |
| D | 1842 | hCoV-19/Spain/PV-IBV-001628/2020\|EPI_ISL_467149\|20 | ···················· | ···················· | ···················· | ···················· | ···················· | ···················· | |
| D | 1843 | hCoV-19/Spain/PV-IBV-001499/2020\|EPI_ISL_467150\|20 | ···················· | ···················· | ···················· | ···················· | ···················· | ···················· | |
| D | 1844 | hCoV-19/Spain/PV-IBV-001561/2020\|EPI_ISL_467151\|20 | ···················· | ·A·················· | ···················· | ···················· | ·NNN················ | ···················· | 1 |
| D | 1845 | hCoV-19/Spain/PV-IBV-001500/2020\|EPI_ISL_467152\|20 | ···················· | ···················· | ···················· | ···················· | ···················· | ···················· | |
| D | 1846 | hCoV-19/Spain/PV-IBV-001612/2020\|EPI_ISL_467153\|20 | ···················· | ···················· | ···················· | ···················· | ···················· | ···················· | |
| D | 1847 | hCoV-19/Spain/PV-IBV-001452/2020\|EPI_ISL_467154\|20 | ···················· | ···················· | ···················· | ···················· | ···················· | ···················· | |
| D | 1848 | hCoV-19/Spain/PV-IBV-001470/2020\|EPI_ISL_467155\|20 | ···················· | ···················· | ···················· | ···················· | ···················· | ···················· | |
| D | 1849 | hCoV-19/Spain/PV-IBV-001577/2020\|EPI_ISL_467156\|20 | ···················· | ···················· | ···················· | ···················· | ···················· | ···················· | |
| D | 1850 | hCoV-19/Spain/PV-IBV-001498/2020\|EPI_ISL_467157\|20 | ···················· | ···················· | ···················· | ···················· | ···················· | ···················· | |
| D | 1851 | hCoV-19/Spain/PV-IBV-001610/2020\|EPI_ISL_467158\|20 | ···················· | ···················· | ···················· | ···················· | ···················· | ···················· | |
| D | 1852 | hCoV-19/Spain/PV-IBV-001528/2020\|EPI_ISL_467159\|20 | ···················· | ···················· | ···················· | ···················· | ···················· | ···················· | |
| D | 1853 | hCoV-19/Spain/PV-IBV-001598/2020\|EPI_ISL_467160\|20 | ···················· | ···················· | ···················· | ···················· | ···················· | ···················· | |
| D | 1854 | hCoV-19/Spain/PV-IBV-001608/2020\|EPI_ISL_467161\|20 | ···················· | ···················· | ···················· | ···················· | ···················· | ···················· | |
| D | 1855 | hCoV-19/Spain/PV-IBV-001485/2020\|EPI_ISL_467162\|20 | ···················· | ···················· | ···················· | ···················· | ···················· | ···················· | |
| D | 1856 | hCoV-19/Spain/PV-IBV-001478/2020\|EPI_ISL_467163\|20 | ···················· | ···················· | ···················· | ···················· | ···················· | ···················· | |
| D | 1857 | hCoV-19/Spain/PV-IBV-001605/2020\|EPI_ISL_467164\|20 | ···················· | ···················· | ···················· | ···················· | ···················· | ···················· | |
| D | 1858 | hCoV-19/Spain/PV-IBV-001606/2020\|EPI_ISL_467165\|20 | ···················· | ···················· | ···················· | ···················· | ···················· | ···················· | |
| D | 1859 | hCoV-19/Spain/PV-IBV-001484/2020\|EPI_ISL_467166\|20 | ···················· | ···················· | ···················· | ···················· | ···················· | ···················· | |
| D | 1860 | hCoV-19/Spain/PV-IBV-001525/2020\|EPI_ISL_467167\|20 | ···················· | ·A·················· | ···················· | ···················· | ···················· | ···················· | |
| D | 1861 | hCoV-19/Spain/PV-IBV-001616/2020\|EPI_ISL_467168\|20 | ···················· | ···················· | ···················· | ···················· | ···················· | ···················· | |
| D | 1862 | hCoV-19/Spain/PV-IBV-001621/2020\|EPI_ISL_467169\|20 | ···················· | ···················· | ···················· | ···················· | ···················· | ···················· | |
| D | 1863 | hCoV-19/USA/CA-CZB-1452/2020\|EPI_ISL_468415\|2020-0 | ···················· | ···················· | ···················· | ···················· | ···················· | ···················· | |
| D | 1864 | hCoV-19/USA/CA-CZB-1456/2020\|EPI_ISL_468417\|2020-0 | ···················· | ···················· | ···················· | ···················· | ···················· | ···················· | |
| D | 1865 | hCoV-19/USA/CA-CZB-1457/2020\|EPI_ISL_468418\|2020-0 | ···················· | ···················· | ···················· | ···················· | ···················· | ···················· | |
| D | 1866 | hCoV-19/USA/CA-CZB-1458/2020\|EPI_ISL_468419\|2020-0 | ···················· | ···················· | ···················· | ···················· | ···················· | ···················· | |
| D | 1867 | hCoV-19/USA/CA-CZB-1459/2020\|EPI_ISL_468420\|2020-0 | ···················· | ···················· | ···················· | ···················· | ···················· | ···················· | |
| D | 1868 | hCoV-19/USA/CA-CZB-1460/2020\|EPI_ISL_468421\|2020-0 | ···················· | ···················· | ···················· | ···················· | ···················· | ···················· | |
| D | 1869 | hCoV-19/USA/CA-CZB-1461/2020\|EPI_ISL_468422\|2020-0 | ···················· | ···················· | ···················· | ···················· | ···················· | ···················· | |
| D | 1870 | hCoV-19/USA/CA-CZB-1464/2020\|EPI_ISL_468424\|2020-0 | ···················· | ···················· | ···················· | ···················· | ···················· | ···················· | |
| D | 1871 | hCoV-19/USA/CA-CZB-1466/2020\|EPI_ISL_468425\|2020-0 | ···················· | ···················· | ···················· | ···················· | ···················· | ···················· | |
| D | 1872 | hCoV-19/USA/CA-CZB-1469/2020\|EPI_ISL_468426\|2020-0 | ···················· | ···················· | ···················· | ···················· | ···················· | ···················· | |
| D | 1873 | hCoV-19/USA/CA-CZB-1473/2020\|EPI_ISL_468427\|2020-0 | ···················· | ···················· | ···················· | ···················· | ···················· | ···················· | |
| D | 1874 | hCoV-19/USA/CA-CZB-1477/2020\|EPI_ISL_468429\|2020-0 | ···················· | ···················· | ···················· | ···················· | ···················· | ···················· | |
| D | 1875 | hCoV-19/USA/CA-CZB-1478/2020\|EPI_ISL_468430\|2020-0 | ···················· | ···················· | ···················· | ···················· | ···················· | ···················· | |
| D | 1876 | hCoV-19/USA/CA-CZB-1479/2020\|EPI_ISL_468431\|2020-0 | ···················· | ···················· | ···················· | ···················· | ···················· | ···················· | |
| D | 1877 | hCoV-19/USA/CA-CZB-1481/2020\|EPI_ISL_468432\|2020-0 | ···················· | ···················· | ···················· | ···················· | ···················· | ···················· | |
| D | 1878 | hCoV-19/USA/CA-CZB-1482/2020\|EPI_ISL_468433\|2020-0 | ···················· | ···················· | ···················· | ···················· | ···················· | ···················· | |
| D | 1879 | hCoV-19/USA/CA-CZB-1484/2020\|EPI_ISL_468435\|2020-0 | ···················· | ···················· | ···················· | ···················· | ···················· | ···················· | |
| D | 1880 | hCoV-19/USA/CA-CZB-1485/2020\|EPI_ISL_468436\|2020-0 | ···················· | ···················· | ···················· | ···················· | ···················· | ···················· | |
| D | 1881 | hCoV-19/USA/CA-CZB-1486/2020\|EPI_ISL_468437\|2020-0 | ···················· | ···················· | ···················· | ···················· | ···················· | ···················· | |
| D | 1882 | hCoV-19/USA/CA-CZB-1488/2020\|EPI_ISL_468438\|2020-0 | ···················· | ···················· | ···················· | ···················· | ···················· | ···················· | |
| D | 1883 | hCoV-19/USA/CA-CZB-1490/2020\|EPI_ISL_468440\|2020-0 | ···················· | ···················· | ···················· | ···················· | ···················· | ···················· | |
| D | 1884 | hCoV-19/USA/CA-CZB-1497/2020\|EPI_ISL_468446\|2020-0 | ···················· | ···················· | ···················· | ···················· | ···················· | ···················· | |
| D | 1885 | hCoV-19/USA/CA-CZB-1498/2020\|EPI_ISL_468447\|2020-0 | ···················· | ···················· | ···················· | ···················· | ···················· | ···················· | |
| D | 1886 | hCoV-19/USA/CA-CZB-1499/2020\|EPI_ISL_468448\|2020-0 | ···················· | ···················· | ···················· | ···················· | ···················· | ···················· | |
| D | 1887 | hCoV-19/USA/CA-CZB-1502/2020\|EPI_ISL_468449\|2020-0 | ···················· | ···················· | ···················· | ···················· | ···················· | ···················· | |
| D | 1888 | hCoV-19/USA/CA-CZB-1503/2020\|EPI_ISL_468450\|2020-0 | ···················· | ···················· | ···················· | ···················· | ···················· | ···················· | |
| D | 1889 | hCoV-19/USA/CA-CZB-1505/2020\|EPI_ISL_468452\|2020-0 | ···················· | ···················· | ···················· | ···················· | ···················· | ···················· | |
| D | 1890 | hCoV-19/USA/CA-CZB-1506/2020\|EPI_ISL_468453\|2020-0 | ···················· | ···················· | ···················· | ···················· | ···················· | ···················· | |
| D | 1891 | hCoV-19/USA/CA-CZB-1509/2020\|EPI_ISL_468456\|2020-0 | ···················· | ···················· | ···················· | ···················· | ···················· | ···················· | |
| D | 1892 | hCoV-19/USA/CA-CZB-1511/2020\|EPI_ISL_468458\|2020-0 | ···················· | ···················· | ···················· | ···················· | ···················· | ···················· | |
| D | 1893 | hCoV-19/USA/CA-CZB-1524/2020\|EPI_ISL_468470\|2020-0 | ···················· | ···················· | ···················· | ···················· | ···················· | ···················· | |
| D | 1894 | hCoV-19/USA/CA-CZB-1528/2020\|EPI_ISL_468473\|2020-0 | ···················· | ···················· | ···················· | ···················· | ···················· | ···················· | |
| D | 1895 | hCoV-19/USA/CA-CZB-1530/2020\|EPI_ISL_468475\|2020-0 | ···················· | ·A·················· | ···················· | ···················· | ···················· | ···················· | |
| D | 1896 | hCoV-19/USA/CA-CZB-1531/2020\|EPI_ISL_468476\|2020-0 | ···················· | ···················· | ···················· | ···················· | ···················· | ···················· | |
| D | 1897 | hCoV-19/USA/CA-CZB-1532/2020\|EPI_ISL_468477\|2020-0 | ···················· | ···················· | ···················· | ···················· | ···················· | ···················· | |
| D | 1898 | hCoV-19/USA/CA-CZB-1533/2020\|EPI_ISL_468478\|2020-0 | ···················· | ·A·················· | ···················· | ···················· | ···················· | ···················· | |
| D | 1899 | hCoV-19/USA/CA-CZB-1534/2020\|EPI_ISL_468479\|2020-0 | ···················· | ···················· | ···················· | ···················· | ···················· | ···················· | |
| D | 1900 | hCoV-19/USA/CA-CZB-1535/2020\|EPI_ISL_468480\|2020-0 | ···················· | ···················· | ···················· | ···················· | ···················· | ···················· | |
| D | 1901 | hCoV-19/USA/CA-CZB-1536/2020\|EPI_ISL_468481\|2020-0 | ···················· | ···················· | ···················· | ···················· | ···················· | ···················· | |
| D | 1902 | hCoV-19/USA/CA-CZB-1537/2020\|EPI_ISL_468482\|2020-0 | ···················· | ···················· | ···················· | ···················· | ···················· | ···················· | |

| | | | | | | | |
|---|---|---|---|---|---|---|---|
| D | 1903 hCoV-19/USA/CA-CZB-1538/2020\|EPI_ISL_468483\|2020-0 | ···················· | ···················· | ···················· | ···················· | ···················· | ···················· |
| D | 1904 hCoV-19/USA/CA-CZB-1539/2020\|EPI_ISL_468484\|2020-0 | ···················· | ···················· | ···················· | ···················· | ···················· | ···················· |
| D | 1905 hCoV-19/USA/CA-CZB-1540/2020\|EPI_ISL_468485\|2020-0 | ···················· | ···················· | ···················· | ···················· | ···················· | ···················· |
| D | 1906 hCoV-19/USA/CA-CZB-1541/2020\|EPI_ISL_468486\|2020-0 | ···················· | ···················· | ···················· | ···················· | ···················· | ···················· |
| D | 1907 hCoV-19/USA/CA-CZB-1542/2020\|EPI_ISL_468487\|2020-0 | ···················· | ···················· | ···················· | ···················· | ···················· | ···················· |
| D | 1908 hCoV-19/USA/CA-CZB-1544/2020\|EPI_ISL_468489\|2020-0 | ···················· | ···················· | ···················· | ···················· | ···················· | ···················· |
| D | 1909 hCoV-19/USA/CA-CZB-1545/2020\|EPI_ISL_468490\|2020-0 | ···················· | ···················· | ···················· | ···················· | ···················· | ···················· |
| D | 1910 hCoV-19/USA/CA-CZB-1546/2020\|EPI_ISL_468491\|2020-0 | ···················· | ···················· | ···················· | ···················· | ···················· | ···················· |
| D | 1911 hCoV-19/USA/CA-CZB-1547/2020\|EPI_ISL_468492\|2020-0 | ···················· | ···················· | ···················· | ···················· | ···················· | ···················· |
| D | 1912 hCoV-19/USA/CA-CZB-1549/2020\|EPI_ISL_468493\|2020-0 | ···················· | ···················· | ···················· | ···················· | ···················· | ···················· |
| D | 1913 hCoV-19/USA/CA-CZB-1551/2020\|EPI_ISL_468494\|2020-0 | ···················· | ···················· | ···················· | ···················· | ···················· | ···················· |
| D | 1914 hCoV-19/USA/CA-CZB-1555/2020\|EPI_ISL_468495\|2020-0 | ···················· | ···················· | ···················· | ···················· | ···················· | ···················· |
| D | 1915 hCoV-19/USA/CA-CZB-1559/2020\|EPI_ISL_468496\|2020-0 | ···················· | ···················· | ···················· | ···················· | ···················· | ···················· |
| D | 1916 hCoV-19/USA/CA-CZB-1560/2020\|EPI_ISL_468497\|2020-0 | ···················· | ···················· | ···················· | ···················· | ···················· | ···················· |
| D | 1917 hCoV-19/USA/CA-CZB-1561/2020\|EPI_ISL_468498\|2020-0 | ···················· | ···················· | ···················· | ···················· | ···················· | ···················· |
| D | 1918 hCoV-19/USA/CA-CZB-1562/2020\|EPI_ISL_468499\|2020-0 | ···················· | ···················· | ···················· | ···················· | ···················· | ···················· |
| D | 1919 hCoV-19/England/BRIS-127E44/2020\|EPI_ISL_469746\|20 | ···················· | ···················· | ···················· | ···················· | ···················· | ···················· |
| D | 1920 hCoV-19/England/BRIS-128344/2020\|EPI_ISL_469747\|20 | ···················· | ···················· | ···················· | ···················· | ···················· | ···················· |
| D | 1921 hCoV-19/England/BRIS-127688/2020\|EPI_ISL_469749\|20 | ···················· | ···················· | ···················· | ···················· | ···················· | ···················· |
| D | 1922 hCoV-19/England/BRIS-127442/2020\|EPI_ISL_469750\|20 | ···················· | ···················· | ···················· | ···················· | ···················· | ···················· |
| D | 1923 hCoV-19/England/BRIS-127961/2020\|EPI_ISL_469752\|20 | ···················· | ···················· | ···················· | ···················· | ···················· | ···················· |
| D | 1924 hCoV-19/England/BRIS-128238/2020\|EPI_ISL_469753\|20 | ···················· | ···················· | ···················· | ···················· | ···················· | ···················· |
| D | 1925 hCoV-19/England/BRIS-126F15/2020\|EPI_ISL_469755\|20 | ···················· | ····A··············· | ···················· | ···················· | ···················· | ···················· |
| D | 1926 hCoV-19/England/BRIS-12755E/2020\|EPI_ISL_469758\|20 | ···················· | ···················· | ···················· | ···················· | ···················· | ···················· |
| D | 1927 hCoV-19/England/BRIS-127C86/2020\|EPI_ISL_469760\|20 | ···················· | ···················· | ···················· | ···················· | ···················· | ···················· |
| D | 1928 hCoV-19/England/BRIS-127D56/2020\|EPI_ISL_469763\|20 | ···················· | ···················· | ···················· | ···················· | ···················· | ···················· |
| D | 1929 hCoV-19/England/BRIS-1274D9/2020\|EPI_ISL_469765\|20 | ···················· | ···················· | ···················· | ···················· | ···················· | ···················· |
| D | 1930 hCoV-19/England/BRIS-1279F8/2020\|EPI_ISL_469766\|20 | ···················· | ···················· | ···················· | ···················· | ···················· | ···················· |
| D | 1931 hCoV-19/England/BRIS-127336/2020\|EPI_ISL_469767\|20 | ···················· | ···················· | ···················· | ···················· | ···················· | ···················· |
| D | 1932 hCoV-19/England/BRIS-12854E/2020\|EPI_ISL_469768\|20 | ···················· | ···················· | ···················· | ···················· | ···················· | ···················· |
| D | 1933 hCoV-19/England/BRIS-1277B2/2020\|EPI_ISL_469771\|20 | ···················· | ···················· | ···················· | ···················· | ···················· | ···················· |
| D | 1934 hCoV-19/England/BRIS-127BE3/2020\|EPI_ISL_469772\|20 | ···················· | ···················· | ···················· | ···················· | ···················· | ···················· |
| D | 1935 hCoV-19/England/BRIS-1282B0/2020\|EPI_ISL_469773\|20 | ···················· | ···················· | ···················· | ···················· | ···················· | ···················· |
| D | 1936 hCoV-19/England/BRIS-127BB6/2020\|EPI_ISL_469774\|20 | ···················· | ···················· | ···················· | ···················· | ···················· | ···················· |
| D | 1937 hCoV-19/England/BRIS-1281FF/2020\|EPI_ISL_469775\|20 | ···················· | ···················· | ···················· | ···················· | ···················· | ···················· |
| D | 1938 hCoV-19/England/BRIS-126F9D/2020\|EPI_ISL_469777\|20 | ···················· | ···················· | ···················· | ···················· | ···················· | ···················· |
| D | 1939 hCoV-19/England/BRIS-127381/2020\|EPI_ISL_469779\|20 | ···················· | ···················· | ···················· | ···················· | ···················· | ···················· |
| D | 1940 hCoV-19/England/BRIS-127C77/2020\|EPI_ISL_469781\|20 | ···················· | ···················· | ···················· | ···················· | ···················· | ···················· |
| D | 1941 hCoV-19/England/BRIS-127D47/2020\|EPI_ISL_469782\|20 | ···················· | ···················· | ···················· | ···················· | ···················· | ···················· |
| D | 1942 hCoV-19/England/BRIS-127169/2020\|EPI_ISL_469785\|20 | ···················· | ···················· | ···················· | ···················· | ···················· | ···················· |
| D | 1943 hCoV-19/England/BRIS-128256/2020\|EPI_ISL_469786\|20 | ···················· | ···················· | ···················· | ···················· | ···················· | ···················· |
| D | 1944 hCoV-19/England/BRIS-128362/2020\|EPI_ISL_469787\|20 | ···················· | ···················· | ···················· | ···················· | ···················· | ···················· |
| D | 1945 hCoV-19/England/BRIS-127011/2020\|EPI_ISL_469788\|20 | ···················· | ····A··············· | ···················· | ···················· | ···················· | ···················· |
| D | 1946 hCoV-19/England/BRIS-127FBA/2020\|EPI_ISL_469789\|20 | ···················· | ···················· | ···················· | ···················· | ···················· | ···················· |
| D | 1947 hCoV-19/USA/UT-02130/2020\|EPI_ISL_470732\|2020-04-1 | ···················· | ···················· | ···················· | ···················· | ···················· | ···················· |
| D | 1948 hCoV-19/USA/UT-02132/2020\|EPI_ISL_470734\|2020-04-1 | ···················· | ···················· | ···················· | ···················· | ···················· | ···················· |
| D | 1949 hCoV-19/USA/UT-03173/2020\|EPI_ISL_470738\|2020-04-2 | ···················· | ···················· | ···················· | ···················· | ···················· | ···················· |
| D | 1950 hCoV-19/USA/UT-03180/2020\|EPI_ISL_470740\|2020-04-2 | ···················· | ···················· | ···················· | ···················· | ···················· | ···················· |
| D | 1951 hCoV-19/USA/UT-03186/2020\|EPI_ISL_470741\|2020-04-2 | ···················· | ···················· | ···················· | ···················· | ···················· | ···················· |
| D | 1952 hCoV-19/USA/UT-01888/2020\|EPI_ISL_470743\|2020-04-1 | ···················· | ···················· | ···················· | ···················· | ···················· | ···················· |
| D | 1953 hCoV-19/USA/UT-01891/2020\|EPI_ISL_470745\|2020-04-1 | ···················· | ···················· | ···················· | ···················· | ···················· | ···················· |
| D | 1954 hCoV-19/USA/MN-MDH-1067/2020\|EPI_ISL_470747\|2020-0 | ···················· | ·A·················· | ···················· | ···················· | ···················· | ···················· |
| D | 1955 hCoV-19/USA/MN-MDH-1068/2020\|EPI_ISL_470748\|2020-0 | ···················· | ·A·················· | ···················· | ···················· | ···················· | ···················· |
| D | 1956 hCoV-19/USA/MN-MDH-1069/2020\|EPI_ISL_470749\|2020-0 | ···················· | ·A·················· | ···················· | ···················· | ···················· | ···················· |
| D | 1957 hCoV-19/USA/MN-MDH-1070/2020\|EPI_ISL_470750\|2020-0 | ···················· | ···················· | ···················· | ···················· | ···················· | ···················· |
| D | 1958 hCoV-19/USA/MN-MDH-1071/2020\|EPI_ISL_470751\|2020-0 | ···················· | ···················· | ···················· | ···················· | ···················· | ···················· |
| D | 1959 hCoV-19/USA/MN-MDH-1072/2020\|EPI_ISL_470752\|2020-0 | ···················· | ···················· | ···················· | ···················· | ···················· | ···················· |
| D | 1960 hCoV-19/USA/MN-MDH-1073/2020\|EPI_ISL_470753\|2020-0 | ···················· | ···················· | ···················· | ···················· | ···················· | ···················· |
| D | 1961 hCoV-19/USA/MN-MDH-1074/2020\|EPI_ISL_470754\|2020-0 | ···················· | ···················· | ···················· | ···················· | ···················· | ···················· |
| D | 1962 hCoV-19/USA/MN-MDH-1075/2020\|EPI_ISL_470755\|2020-0 | ···················· | ···················· | ···················· | ···················· | ···················· | ···················· |
| D | 1963 hCoV-19/USA/MN-MDH-1076/2020\|EPI_ISL_470756\|2020-0 | ···················· | ···················· | ···················· | ···················· | ···················· | ···················· |
| D | 1964 hCoV-19/USA/MN-MDH-1079/2020\|EPI_ISL_470759\|2020-0 | ···················· | ···················· | ···················· | ···················· | ···················· | ···················· |
| D | 1965 hCoV-19/USA/MN-MDH-1090/2020\|EPI_ISL_470770\|2020-0 | ···················· | ···················· | ···················· | ···················· | ···················· | ···················· |
| D | 1966 hCoV-19/USA/MN-MDH-1091/2020\|EPI_ISL_470771\|2020-0 | ···················· | ···················· | ···················· | ···················· | ···················· | ···················· |
| D | 1967 hCoV-19/USA/MN-MDH-1092/2020\|EPI_ISL_470772\|2020-0 | ···················· | ···················· | ···················· | ···················· | ···················· | ···················· |
| D | 1968 hCoV-19/USA/MN-MDH-1093/2020\|EPI_ISL_470773\|2020-0 | ···················· | ···················· | ···················· | ···················· | ···················· | ···················· |
| D | 1969 hCoV-19/USA/MN-MDH-1094/2020\|EPI_ISL_470774\|2020-0 | ···················· | ···················· | ···················· | ···················· | ···················· | ···················· |
| D | 1970 hCoV-19/USA/MN-MDH-1095/2020\|EPI_ISL_470775\|2020-0 | ···················· | ···················· | ···················· | ···················· | ···················· | ···················· |
| D | 1971 hCoV-19/USA/MN-MDH-1096/2020\|EPI_ISL_470776\|2020-0 | ···················· | ···················· | ···················· | ···················· | ···················· | ···················· |
| D | 1972 hCoV-19/USA/MN-MDH-1097/2020\|EPI_ISL_470777\|2020-0 | ···················· | ···················· | ···················· | ···················· | ···················· | ···················· |
| D | 1973 hCoV-19/USA/MN-MDH-1098/2020\|EPI_ISL_470778\|2020-0 | ···················· | ···················· | ···················· | ···················· | ···················· | ···················· |
| D | 1974 hCoV-19/USA/MN-MDH-1099/2020\|EPI_ISL_470779\|2020-0 | ···················· | ···················· | ···················· | ···················· | ···················· | ···················· |
| D | 1975 hCoV-19/USA/MN-MDH-1100/2020\|EPI_ISL_470780\|2020-0 | ···················· | ···················· | ···················· | ···················· | ···················· | ···················· |
| D | 1976 hCoV-19/USA/MN-MDH-1101/2020\|EPI_ISL_470781\|2020-0 | ···················· | ···················· | ···················· | ···················· | ···················· | ···················· |
| D | 1977 hCoV-19/USA/MN-MDH-1102/2020\|EPI_ISL_470782\|2020-0 | ···················· | ···················· | ···················· | ···················· | ···················· | ···················· |
| D | 1978 hCoV-19/USA/MN-MDH-1103/2020\|EPI_ISL_470783\|2020-0 | ···················· | ···················· | ···················· | ···················· | ···················· | ···················· |
| D | 1979 hCoV-19/USA/MN-MDH-1104/2020\|EPI_ISL_470784\|2020-0 | ···················· | ···················· | ···················· | ···················· | ···················· | ···················· |
| D | 1980 hCoV-19/USA/MN-MDH-1105/2020\|EPI_ISL_470785\|2020-0 | ···················· | ···················· | ···················· | ···················· | ···················· | ···················· |

| | | | | | | | | |
|---|---|---|---|---|---|---|---|---|
| D | 1981 hCoV-19/USA/MN-MDH-1106/2020\|EPI_ISL_470786\|2020-0 | ···················· | ···················· | ···················· | ···················· | ···················· | ···················· | |
| D | 1982 hCoV-19/USA/MN-MDH-1107/2020\|EPI_ISL_470787\|2020-0 | ···················· | ···················· | ···················· | ···················· | ···················· | ···················· | |
| D | 1983 hCoV-19/USA/MN-MDH-1108/2020\|EPI_ISL_470788\|2020-0 | ···················· | ···················· | ···················· | ···················· | ···················· | ···················· | |
| D | 1984 hCoV-19/USA/MN-MDH-1109/2020\|EPI_ISL_470789\|2020-0 | ···················· | ···················· | ···················· | ···················· | ···················· | ···················· | |
| D | 1985 hCoV-19/USA/MN-MDH-1110/2020\|EPI_ISL_470790\|2020-0 | ···················· | ·A·················· | ···················· | ···················· | ···················· | ···················· | |
| D | 1986 hCoV-19/USA/MN-MDH-1111/2020\|EPI_ISL_470791\|2020-0 | ···················· | ·A·················· | ···················· | ···················· | ···················· | ···················· | |
| D | 1987 hCoV-19/USA/MN-MDH-1112/2020\|EPI_ISL_470792\|2020-0 | ···················· | ·A·················· | ···················· | ···················· | ···················· | ···················· | |
| D | 1988 hCoV-19/USA/MN-MDH-1113/2020\|EPI_ISL_470793\|2020-0 | ···················· | ·A·················· | ···················· | ···················· | ···················· | ···················· | |
| D | 1989 hCoV-19/USA/MN-MDH-1114/2020\|EPI_ISL_470794\|2020-0 | ···················· | ·A·················· | ···················· | ···················· | ···················· | ···················· | |
| D | 1990 hCoV-19/USA/MN-MDH-1115/2020\|EPI_ISL_470795\|2020-0 | ···················· | ·A·················· | ···················· | ···················· | ···················· | ···················· | |
| D | 1991 hCoV-19/USA/MN-MDH-1116/2020\|EPI_ISL_470796\|2020-0 | ···················· | ·A·················· | ···················· | ···················· | ···················· | ···················· | |
| D | 1992 hCoV-19/USA/MN-MDH-1117/2020\|EPI_ISL_470797\|2020-0 | ···················· | ·A·················· | ···················· | ···················· | ···················· | ···················· | |
| D | 1993 hCoV-19/USA/MN-MDH-1118/2020\|EPI_ISL_470798\|2020-0 | ···················· | ·A·················· | ···················· | ···················· | ···················· | ···················· | |
| D | 1994 hCoV-19/USA/MN-MDH-1119/2020\|EPI_ISL_470799\|2020-0 | ···················· | ·A·················· | ···················· | ···················· | ···················· | ···················· | |
| D | 1995 hCoV-19/USA/WA-UW-4755/2020\|EPI_ISL_476900\|2020-03 | ···················· | ···················· | ···················· | ···················· | ···················· | ···················· | |
| D | 1996 hCoV-19/USA/WA-UW-1128/2020\|EPI_ISL_476902\|2020-03 | ···················· | ···················· | ···················· | ···················· | ···················· | ···················· | |
| D | 1997 hCoV-19/USA/WA-UW-114/2020\|EPI_ISL_476903\|2020-03- | ···················· | ···················· | ···················· | ···················· | ···················· | ···················· | |
| D | 1998 hCoV-19/USA/WA-UW-1274/2020\|EPI_ISL_476905\|2020-03 | ···················· | ···················· | ···················· | ···················· | ···················· | ···················· | |
| D | 1999 hCoV-19/USA/WA-UW-571/2020\|EPI_ISL_476906\|2020-03- | ···················· | ···················· | ···················· | ···················· | ···················· | ···················· | |
| D | 2000 hCoV-19/USA/WA-UW-608/2020\|EPI_ISL_476907\|2020-03- | ···················· | ···················· | ···················· | ···················· | ···················· | ···················· | |
| D | 2001 hCoV-19/USA/WA-UW-623/2020\|EPI_ISL_476908\|2020-03- | ···················· | ···················· | ···················· | ···················· | ···················· | ···················· | |
| D | 2002 hCoV-19/USA/MN-UW-628/2020\|EPI_ISL_476909\|2020-03- | ···················· | ···················· | ···················· | ···················· | ···················· | ···················· | |
| D | 2003 hCoV-19/England/OXON-B203A/2020\|EPI_ISL_479107\|202 | ···················· | ···················· | ···················· | ···················· | ···················· | ···················· | |
| D | 2004 hCoV-19/England/OXON-B2049/2020\|EPI_ISL_479108\|202 | ···················· | ···················· | ···················· | ···················· | ···················· | ···················· | |
| D | 2005 hCoV-19/England/OXON-B2076/2020\|EPI_ISL_479110\|202 | ···················· | ···················· | ···················· | ···················· | ···················· | ···················· | |
| D | 2006 hCoV-19/England/OXON-B2085/2020\|EPI_ISL_479111\|202 | ···················· | ···················· | ···················· | ···················· | ···················· | ···················· | |
| D | 2007 hCoV-19/England/OXON-B20C1/2020\|EPI_ISL_479113\|202 | ···················· | ···················· | ···················· | ···················· | ···················· | ···················· | |
| D | 2008 hCoV-19/England/OXON-B20EF/2020\|EPI_ISL_479114\|202 | ···················· | ···················· | ···················· | ···················· | ···················· | ···················· | |
| D | 2009 hCoV-19/England/OXON-B20FE/2020\|EPI_ISL_479115\|202 | ···················· | ···················· | ···················· | ···················· | ···················· | ···················· | |
| D | 2010 hCoV-19/England/OXON-B210A/2020\|EPI_ISL_479116\|202 | ···················· | ···················· | ···················· | ···················· | ···················· | ···················· | |
| D | 2011 hCoV-19/England/OXON-B2119/2020\|EPI_ISL_479117\|202 | ···················· | ···················A | ···················· | ···················· | ···················· | ···················· | 1 |
| D | 2012 hCoV-19/England/OXON-B2137/2020\|EPI_ISL_479119\|202 | ···················· | ···················· | ···················· | ···················· | ···················· | ···················· | |
| D | 2013 hCoV-19/England/OXON-B2146/2020\|EPI_ISL_479120\|202 | ···················· | ···················· | ···················· | ···················· | ···················· | ···················· | |
| D | 2014 hCoV-19/England/OXON-B2155/2020\|EPI_ISL_479121\|202 | ···················· | ···················· | ···················· | ···················· | ···················· | ···················· | |
| D | 2015 hCoV-19/England/OXON-B2164/2020\|EPI_ISL_479122\|202 | ···················· | ···················· | ···················· | ···················· | ···················· | ···················· | |
| D | 2016 hCoV-19/England/OXON-B2173/2020\|EPI_ISL_479123\|202 | ···················· | ···················· | ···················· | ···················· | ···················· | ···················· | |
| D | 2017 hCoV-19/England/OXON-B2191/2020\|EPI_ISL_479124\|202 | ···················· | ···················· | ···················· | ···················· | ···················· | ···················· | |
| D | 2018 hCoV-19/England/OXON-B21EC/2020\|EPI_ISL_479126\|202 | ···················· | ···················· | ···················· | ···················· | ···················· | ···················· | |
| D | 2019 hCoV-19/England/OXON-B21FB/2020\|EPI_ISL_479127\|202 | ···················· | ···················· | ···················· | ···················· | ···················· | ···················· | |
| D | 2020 hCoV-19/England/OXON-B2234/2020\|EPI_ISL_479130\|202 | ···················· | ···················· | ···················· | ···················· | ···················· | ···················· | |
| D | 2021 hCoV-19/England/OXON-B2252/2020\|EPI_ISL_479131\|202 | ···················· | ···················· | ···················· | ···················· | ···················· | ···················· | |
| D | 2022 hCoV-19/England/OXON-B2261/2020\|EPI_ISL_479132\|202 | ···················· | ···················· | ···················· | ···················· | ···················· | ···················· | |
| D | 2023 hCoV-19/England/OXON-B228F/2020\|EPI_ISL_479133\|202 | ···················· | ···················· | ···················· | ···················· | ···················· | ···················· | |
| D | 2024 hCoV-19/England/OXON-B229E/2020\|EPI_ISL_479134\|202 | ···················· | ···················· | ···················· | ···················· | ···················· | ···················· | |
| D | 2025 hCoV-19/England/OXON-B22BC/2020\|EPI_ISL_479135\|202 | ···················· | ···················· | ···················· | ···················· | ···················· | ···················· | |
| D | 2026 hCoV-19/England/OXON-B2313/2020\|EPI_ISL_479136\|202 | ···················· | ···················· | ···················· | ···················· | ···················· | ···················· | |
| D | 2027 hCoV-19/England/OXON-B2322/2020\|EPI_ISL_479137\|202 | ···················· | ···················· | ···················· | ···················· | ···················· | ···················· | |
| D | 2028 hCoV-19/England/OXON-B236E/2020\|EPI_ISL_479138\|202 | ···················· | ···················· | ···················· | ···················· | ···················· | ···················· | |
| D | 2029 hCoV-19/England/OXON-B23D7/2020\|EPI_ISL_479140\|202 | ···················· | ···················· | ···················· | ···················· | ···················· | ···················· | |
| D | 2030 hCoV-19/England/OXON-B23E6/2020\|EPI_ISL_479141\|202 | ···················· | ···················· | ···················· | ···················· | ···················· | ···················· | |
| D | 2031 hCoV-19/England/OXON-B2401/2020\|EPI_ISL_479142\|202 | ···················· | ···················· | ···················· | ···················· | ···················· | ···················· | |
| D | 2032 hCoV-19/England/OXON-B2410/2020\|EPI_ISL_479143\|202 | ···················· | ···················· | ···················· | ···················· | ···················· | ···················· | |
| D | 2033 hCoV-19/England/OXON-B242F/2020\|EPI_ISL_479144\|202 | ···················· | ·A·················· | ···················· | ···················· | ···················· | ···················· | |
| D | 2034 hCoV-19/England/OXON-B246B/2020\|EPI_ISL_479146\|202 | ···················· | ···················· | ···················· | ···················· | ···················· | ···················· | |
| D | 2035 hCoV-19/England/OXON-B247A/2020\|EPI_ISL_479147\|202 | ···················· | ···················· | ···················· | ···················· | ···················· | ···················· | |
| D | 2036 hCoV-19/England/OXON-B24C5/2020\|EPI_ISL_479148\|202 | ···················· | ···················· | ···················· | ···················· | ···················· | ···················· | |
| D | 2037 hCoV-19/England/OXON-B2753/2020\|EPI_ISL_479174\|202 | ···················· | ···················· | ···················· | ···················· | ···················· | ···················· | |
| D | 2038 hCoV-19/England/PORT-2D12B5/2020\|EPI_ISL_479175\|20 | ···················· | ···················· | ···················· | ···················· | ···················· | ···················· | |
| D | 2039 hCoV-19/England/PORT-2D130D/2020\|EPI_ISL_479180\|20 | ···················· | ···················· | ···················· | ···················· | ···················· | ···················· | |
| D | 2040 hCoV-19/England/PORT-2D131C/2020\|EPI_ISL_479181\|20 | ···················· | ···················· | ···················· | ···················· | ···················· | ···················· | |
| D | 2041 hCoV-19/England/PORT-2D132B/2020\|EPI_ISL_479182\|20 | ···················· | ···················· | ···················· | ···················· | ···················· | ···················· | |
| D | 2042 hCoV-19/England/PORT-2D133A/2020\|EPI_ISL_479183\|20 | ···················· | ···················· | ···················· | ···················· | ···················· | ···················· | |
| D | 2043 hCoV-19/England/PORT-2D1349/2020\|EPI_ISL_479184\|20 | ···················· | ···················· | ···················· | ···················· | ···················· | ···················· | |
| D | 2044 hCoV-19/England/PORT-2D1358/2020\|EPI_ISL_479185\|20 | ···················· | ···················· | ···················· | ···················· | ···················· | ···················· | |
| D | 2045 hCoV-19/England/PORT-2D1367/2020\|EPI_ISL_479186\|20 | ···················· | ···················· | ···················· | ···················· | ···················· | ···················· | |
| D | 2046 hCoV-19/England/PORT-2D1376/2020\|EPI_ISL_479187\|20 | ···················· | ···················· | ···················· | ···················· | ···················· | ···················· | |
| D | 2047 hCoV-19/England/PORT-2D1385/2020\|EPI_ISL_479188\|20 | ···················· | ···················· | ···················· | ···················· | ···················· | ···················· | |
| D | 2048 hCoV-19/England/PORT-2D1394/2020\|EPI_ISL_479189\|20 | ···················· | ···················· | ···················· | ···················· | ···················· | ···················· | |
| D | 2049 hCoV-19/Singapore/313/2020\|EPI_ISL_462431\|2020-03- | ···················· | ···················· | ···················· | ···················· | ···················· | ···················· | |
| D | 2050 hCoV-19/Singapore/314/2020\|EPI_ISL_462432\|2020-03- | ···················· | ···················· | ···················· | ···················· | ···················· | ···················· | |
| D | 2051 hCoV-19/Singapore/315/2020\|EPI_ISL_462433\|2020-02- | ···················· | ···················· | ···················· | ···················· | ···················· | ···················· | |
| D | 2052 hCoV-19/Spain/CT-IrsiCaixa-sp50/2020\|EPI_ISL_46244 | ···················· | ···················· | ···················· | ···················· | ···················· | ···················· | |
| D | 2053 hCoV-19/Spain/CT-IrsiCaixa-sp142/2020\|EPI_ISL_4624 | ···················· | ···················· | ···················· | ···················· | ···················· | ···················· | |
| D | 2054 hCoV-19/Spain/CT-IrsiCaixa-015D0/2020\|EPI_ISL_4624 | ···················· | ···················· | ···················· | ···················· | ···················· | ···················· | |
| D | 2055 hCoV-19/Bosnia and Herzegovina/ChVir7340/2020\|EPI_ | ···················· | ···················· | ···················· | ···················· | ···················· | ···················· | |
| D | 2056 hCoV-19/Bosnia and Herzegovina/ChVir7343/2020\|EPI_ | ···················· | ···················· | ···················· | ···················· | ···················· | ···················· | |
| D | 2057 hCoV-19/Bosnia and Herzegovina/ChVir7344/2020\|EPI_ | ···················· | ···················· | ···················· | ···················· | ···················· | ···················· | |
| D | 2058 hCoV-19/Bosnia and Herzegovina/ChVir7346/2020\|EPI_ | ···················· | ···················· | ···················· | ···················· | ···················· | ···················· | |

| | | | | | | | | | |
|---|---|---|---|---|---|---|---|---|---|
| D | 2059 | hCoV-19/Bosnia and Herzegovina/ChVir7347/2020\|EPI_ | ···················· | ···················· | ···················· | ···················· | ···················· | ···················· | |
| D | 2060 | hCoV-19/Bosnia and Herzegovina/ChVir7348/2020\|EPI_ | ···················· | ···················· | ···················· | ···················· | ···················· | ···················· | |
| D | 2061 | hCoV-19/Bosnia and Herzegovina/ChVir7349/2020\|EPI_ | ···················· | ···················· | ···················· | ···················· | ···················· | ···················· | |
| D | 2062 | hCoV-19/Bosnia and Herzegovina/ChVir7350/2020\|EPI_ | ···················· | ···················· | ···················· | ···················· | ···················· | ···················· | |
| D | 2063 | hCoV-19/Bosnia and Herzegovina/ChVir7352/2020\|EPI_ | ···················· | ···················· | ···················· | ···················· | ···················· | ···················· | |
| D | 2064 | hCoV-19/Bosnia and Herzegovina/ChVir7353/2020\|EPI_ | ···················· | ···················· | ···················· | ···················· | ···················· | ···················· | |
| D | 2065 | hCoV-19/Bosnia and Herzegovina/ChVir7354/2020\|EPI_ | ···················· | ···················· | ···················· | ···················· | ···················· | ···················· | |
| D | 2066 | hCoV-19/Bosnia and Herzegovina/ChVir7387/2020\|EPI_ | ···················· | ···················· | ···················· | ···················· | ···················· | ···················· | |
| D | 2067 | hCoV-19/Bosnia and Herzegovina/ChVir7357/2020\|EPI_ | ···················· | ···················· | ···················· | ···················· | ···················· | ···················· | |
| D | 2068 | hCoV-19/Bosnia and Herzegovina/ChVir7360/2020\|EPI_ | ···················· | ···················· | ···················· | ···················· | ···················· | ···················· | |
| D | 2069 | hCoV-19/Bosnia and Herzegovina/ChVir7361/2020\|EPI_ | ···················· | ···················· | ···················· | ···················· | ···················· | ···················· | |
| D | 2070 | hCoV-19/Bosnia and Herzegovina/ChVir7362/2020\|EPI_ | ···················· | ···················· | ···················· | ···················· | ···················· | ···················· | |
| D | 2071 | hCoV-19/Bosnia and Herzegovina/ChVir7396/2020\|EPI_ | ···················· | ···················· | ···················· | ···················· | ···················· | ···················· | |
| D | 2072 | hCoV-19/Bosnia and Herzegovina/ChVir7365/2020\|EPI_ | ···················· | ···················· | ···················· | ···················· | ···················· | ···················· | |
| D | 2073 | hCoV-19/Bosnia and Herzegovina/ChVir7366/2020\|EPI_ | ···················· | ···················· | ···················· | ···················· | ···················· | ···················· | |
| D | 2074 | hCoV-19/Bosnia and Herzegovina/ChVir7367/2020\|EPI_ | ···················· | ···················· | ···················· | ···················· | ···················· | ···················· | |
| D | 2075 | hCoV-19/Bosnia and Herzegovina/ChVir7402/2020\|EPI_ | ···················· | ···················· | ···················· | ···················· | ···················· | ···················· | |
| D | 2076 | hCoV-19/Bosnia and Herzegovina/ChVir7370/2020\|EPI_ | ···················· | ···················· | ···················· | ···················· | ···················· | ···················· | |
| D | 2077 | hCoV-19/Bosnia and Herzegovina/ChVir7382/2020\|EPI_ | ···················· | ···················· | ···················· | ···················· | ···················· | ···················· | |
| D | 2078 | hCoV-19/Bosnia and Herzegovina/ChVir7372/2020\|EPI_ | ···················· | ···················· | ···················· | ···················· | ···················· | ···················· | |
| D | 2079 | hCoV-19/Bosnia and Herzegovina/ChVir7375/2020\|EPI_ | ···················· | ···················· | ···················· | ···················· | ···················· | ···················· | |
| D | 2080 | hCoV-19/Spain/PV-IBV-001477/2020\|EPI_ISL_467128\|20 | ···················· | ·A·················· | ···················· | ···················· | ···················· | ···················· | |
| D | 2081 | hCoV-19/Spain/PV-IBV-001588/2020\|EPI_ISL_467129\|20 | ···················· | ···················· | ···················· | ···················· | ···················· | ···················· | |
| D | 2082 | hCoV-19/Spain/PV-IBV-001436/2020\|EPI_ISL_467130\|20 | ···················· | ···················· | ···················· | ···················· | ···················· | ···················· | |
| D | 2083 | hCoV-19/Spain/PV-IBV-001546/2020\|EPI_ISL_467131\|20 | ···················· | ···················· | ···················· | ···················· | ···················· | ···················· | |
| D | 2084 | hCoV-19/Spain/PV-IBV-001600/2020\|EPI_ISL_467132\|20 | ···················· | ···················· | ···················· | ···················· | ···················· | ···················· | |
| D | 2085 | hCoV-19/Spain/PV-IBV-001611/2020\|EPI_ISL_467134\|20 | ···················· | ···················· | ···················· | ···················· | ···················· | ···················· | |
| D | 2086 | hCoV-19/Spain/PV-IBV-001585/2020\|EPI_ISL_467136\|20 | ···················· | ···················· | ···················· | ···················· | ···················· | ···················· | |
| D | 2087 | hCoV-19/Spain/PV-IBV-001450/2020\|EPI_ISL_467137\|20 | ···················· | ···················· | ···················· | ···················· | ·NNN················ | ···················· | 1 |
| D | 2088 | hCoV-19/Spain/PV-IBV-001555/2020\|EPI_ISL_467138\|20 | ···················· | ·A·················· | ···················· | ···················· | ···················· | ···················· | |
| D | 2090 | hCoV-19/Spain/PV-IBV-001536/2020\|EPI_ISL_467140\|20 | ···················· | ···················· | ···················· | ···················· | ···················· | ···················· | |
| D | 2092 | hCoV-19/Bosnia and Herzegovina/04-Sarajevo/2020\|EP | ···················· | ···················· | ···················· | ···················· | ···················· | ···················· | |
| D | 2093 | hCoV-19/USA/MO-WUSTL033/2020\|EPI_ISL_467301\|2020-0 | ···················· | ···················· | ···················· | ···················· | ···················· | ···················· | |
| D | 2094 | hCoV-19/USA/MO-WUSTL035/2020\|EPI_ISL_467302\|2020-0 | ···················· | ···················· | ···················· | ···················· | ···················· | ···················· | |
| D | 2095 | hCoV-19/USA/MO-WUSTL055/2020\|EPI_ISL_467304\|2020-0 | ···················· | ···················· | ···················· | ···················· | ···················· | ···················· | |
| D | 2096 | hCoV-19/Canada/BC_09329927/2020\|EPI_ISL_467305\|202 | ···················· | ···················· | ···················· | ···················· | ···················· | ···················· | |
| D | 2097 | hCoV-19/Canada/BC_09452317/2020\|EPI_ISL_467306\|202 | ···················· | ···················· | ···················· | ···················· | ···················· | ···················· | |
| D | 2098 | hCoV-19/Canada/BC_09978786/2020\|EPI_ISL_467309\|202 | ···················· | ···················· | ···················· | ···················· | ···················· | ···················· | |
| D | 2099 | hCoV-19/Canada/BC_10063599/2020\|EPI_ISL_467310\|202 | ···················· | ···················· | ···················· | ···················· | ···················· | ···················· | |
| D | 2100 | hCoV-19/Canada/BC_10111761/2020\|EPI_ISL_467312\|202 | ···················· | ···················· | ···················· | ···················· | ···················· | ···················· | |
| D | 2101 | hCoV-19/Canada/BC_10111910/2020\|EPI_ISL_467313\|202 | ···················· | ···················· | ···················· | ···················· | ···················· | ···················· | |
| D | 2102 | hCoV-19/Canada/BC_10589793/2020\|EPI_ISL_467314\|202 | ···················· | ···················· | ···················· | ···················· | ···················· | ···················· | |
| D | 2103 | hCoV-19/Canada/BC_10592956/2020\|EPI_ISL_467315\|202 | ···················· | ···················· | ···················· | ···················· | ···················· | ···················· | |
| D | 2104 | hCoV-19/Canada/BC_11028219/2020\|EPI_ISL_467317\|202 | ···················· | ···················· | ···················· | ···················· | ···················· | ···················· | |
| D | 2105 | hCoV-19/Canada/BC_11135150/2020\|EPI_ISL_467318\|202 | ···················· | ···················· | ···················· | ···················· | ···················· | ···················· | |
| D | 2106 | hCoV-19/Canada/BC_11184415/2020\|EPI_ISL_467320\|202 | ···················· | ···················· | ···················· | ···················· | ···················· | ···················· | |
| D | 2107 | hCoV-19/Canada/BC_11186723/2020\|EPI_ISL_467321\|202 | ···················· | ···················· | ···················· | ···················· | ···················· | ···················· | |
| D | 2108 | hCoV-19/Canada/BC_11303557/2020\|EPI_ISL_467322\|202 | ···················· | ···················· | ···················· | ···················· | ···················· | ···················· | |
| D | 2109 | hCoV-19/Canada/BC_11637543/2020\|EPI_ISL_467324\|202 | ···················· | ···················· | ···················· | ···················· | ···················· | ···················· | |
| D | 2110 | hCoV-19/USA/CA-CZB-1512/2020\|EPI_ISL_468459\|2020-0 | ···················· | ···················· | ···················· | ···················· | ···················· | ···················· | |
| D | 2111 | hCoV-19/USA/CA-CZB-1514/2020\|EPI_ISL_468461\|2020-0 | ···················· | ···················· | ···················· | ···················· | ···················· | ···················· | |
| D | 2112 | hCoV-19/USA/CA-CZB-1516/2020\|EPI_ISL_468462\|2020-0 | ···················· | ···················· | ···················· | ···················· | ···················· | ···················· | |
| D | 2113 | hCoV-19/USA/CA-CZB-1517/2020\|EPI_ISL_468463\|2020-0 | ···················· | ···················· | ···················· | ···················· | ···················· | ···················· | |
| D | 2114 | hCoV-19/USA/CA-CZB-1518/2020\|EPI_ISL_468464\|2020-0 | ···················· | ···················· | ···················· | ···················· | ···················· | ···················· | |
| D | 2115 | hCoV-19/USA/CA-CZB-1519/2020\|EPI_ISL_468465\|2020-0 | ···················· | ···················· | ···················· | ···················· | ···················· | ···················· | |
| D | 2116 | hCoV-19/USA/CA-CZB-1520/2020\|EPI_ISL_468466\|2020-0 | ···················· | ···················· | ···················· | ···················· | ···················· | ···················· | |
| D | 2117 | hCoV-19/USA/CA-CZB-1521/2020\|EPI_ISL_468467\|2020-0 | ···················· | ···················· | ···················· | ···················· | ···················· | ···················· | |
| D | 2118 | hCoV-19/USA/CA-CZB-1522/2020\|EPI_ISL_468468\|2020-0 | ···················· | ···················· | ···················· | ···················· | ···················· | ···················· | |
| D | 2119 | hCoV-19/USA/CA-CZB-1523/2020\|EPI_ISL_468469\|2020-0 | ···················· | ···················· | ···················· | ···················· | ···················· | ···················· | |
| D | 2120 | hCoV-19/USA/CA-CZB-1525/2020\|EPI_ISL_468471\|2020-0 | ···················· | ···················· | ···················· | ···················· | ···················· | ···················· | |
| D | 2121 | hCoV-19/USA/CA-CZB-1526/2020\|EPI_ISL_468472\|2020-0 | ···················· | ·A·················· | ···················· | ···················· | ···················· | ···················· | |
| D | 2122 | hCoV-19/USA/CA-CZB-1656/2020\|EPI_ISL_468600\|2020-0 | ···················· | ···················· | ···················· | ···················· | ···················· | ···················· | |
| D | 2123 | hCoV-19/USA/CA-CZB-1657/2020\|EPI_ISL_468601\|2020-0 | ···················· | ···················· | ···················· | ···················· | ···················· | ···················· | |
| D | 2124 | hCoV-19/USA/CA-CZB-1658/2020\|EPI_ISL_468602\|2020-0 | ···················· | ···················· | ···················· | ···················· | ···················· | ···················· | |
| D | 2125 | hCoV-19/USA/CA-CZB-1659/2020\|EPI_ISL_468603\|2020-0 | ···················· | ···················· | ···················· | ···················· | ···················· | ···················· | |
| D | 2126 | hCoV-19/USA/CA-CZB-1662/2020\|EPI_ISL_468605\|2020-0 | ···················· | ···················· | ···················· | ···················· | ···················· | ···················· | |
| D | 2127 | hCoV-19/USA/CA-CZB-1664/2020\|EPI_ISL_468607\|2020-0 | ···················· | ···················· | ···················· | ···················· | ···················· | ···················· | |
| D | 2128 | hCoV-19/USA/CA-CZB-1666/2020\|EPI_ISL_468608\|2020-0 | ···················· | ···················· | ···················· | ···················· | ···················· | ···················· | |
| D | 2129 | hCoV-19/USA/CA-CZB-1669/2020\|EPI_ISL_468611\|2020-0 | ···················· | ···················· | ···················· | ···················· | ···················· | ···················· | |
| D | 2130 | hCoV-19/USA/CA-CZB-1670/2020\|EPI_ISL_468612\|2020-0 | ···················· | ···················· | ···················· | ···················· | ···················· | ···················· | |
| D | 2131 | hCoV-19/USA/CA-CZB-1672/2020\|EPI_ISL_468614\|2020-0 | ···················· | ···················· | ···················· | ···················· | ···················· | ···················· | |
| D | 2132 | hCoV-19/USA/CA-CZB-1675/2020\|EPI_ISL_468615\|2020-0 | ···················· | ···················· | ···················· | ···················· | ···················· | ···················· | |
| D | 2133 | hCoV-19/USA/CA-CZB-1676/2020\|EPI_ISL_468616\|2020-0 | ···················· | ···················· | ···················· | ···················· | ···················· | ···················· | |
| D | 2134 | hCoV-19/USA/CA-CZB-1677/2020\|EPI_ISL_468617\|2020-0 | ···················· | ···················· | ···················· | ···················· | ···················· | ···················· | |
| D | 2135 | hCoV-19/USA/CA-CZB-1678/2020\|EPI_ISL_468618\|2020-0 | ···················· | ···················· | ···················· | ···················· | ···················· | ···················· | |
| D | 2136 | hCoV-19/USA/CA-CZB-1683/2020\|EPI_ISL_468621\|2020-0 | ···················· | ···················· | ···················· | ···················· | ···················· | ···················· | |
| D | 2137 | hCoV-19/USA/CA-CZB-1684/2020\|EPI_ISL_468622\|2020-0 | ···················· | ···················· | ···················· | ···················· | ···················· | ···················· | |
| D | 2138 | hCoV-19/USA/CA-CZB-1687/2020\|EPI_ISL_468624\|2020-0 | ···················· | ···················· | ···················· | ···················· | ···················· | ···················· | |

| | | | | | | | | | |
|---|---|---|---|---|---|---|---|---|---|
| D | 2139 | hCoV-19/USA/CA-CZB-1689/2020\|EPI_ISL_468625\|2020-0 | ···················· | ···················· | ···················· | ···················· | ···················· | ···················· | |
| D | 2140 | hCoV-19/USA/CA-CZB-1690/2020\|EPI_ISL_468626\|2020-0 | ···················· | ···················· | ···················· | ···················· | ···················· | ···················· | |
| D | 2141 | hCoV-19/USA/CA-CZB-1691/2020\|EPI_ISL_468627\|2020-0 | ···················· | ···················· | ···················· | ···················· | ···················· | ···················· | |
| D | 2142 | hCoV-19/USA/CA-CZB-1693/2020\|EPI_ISL_468628\|2020-0 | ···················· | ···················· | ···················· | ···················· | ···················· | ···················· | |
| D | 2143 | hCoV-19/USA/CA-CZB-1694/2020\|EPI_ISL_468629\|2020-0 | ···················· | ···················· | ···················· | ···················· | ···················· | ···················· | |
| D | 2144 | hCoV-19/USA/CA-CZB-1696/2020\|EPI_ISL_468631\|2020-0 | ···················· | ···················· | ···················· | ···················· | ···················· | ···················· | |
| D | 2145 | hCoV-19/USA/CA-CZB-1697/2020\|EPI_ISL_468632\|2020-0 | ···················· | ···················· | ···················· | ···················· | ···················· | ···················· | |
| D | 2146 | hCoV-19/USA/CA-CZB-1698/2020\|EPI_ISL_468633\|2020-0 | ···················· | ···················· | ···················· | ···················· | ···················· | ···················· | |
| D | 2147 | hCoV-19/USA/CA-CZB-1700/2020\|EPI_ISL_468634\|2020-0 | ···················· | ···················· | ···················· | ···················· | ···················· | ···················· | |
| D | 2148 | hCoV-19/USA/CA-CZB-1702/2020\|EPI_ISL_468636\|2020-0 | ···················· | ···················· | ···················· | ···················· | ···················· | ···················· | |
| D | 2149 | hCoV-19/USA/CA-CZB-1704/2020\|EPI_ISL_468637\|2020-0 | ···················· | ···················· | ···················· | ···················· | ···················· | ···················· | |
| D | 2150 | hCoV-19/USA/CA-CZB-1705/2020\|EPI_ISL_468638\|2020-0 | ···················· | ···················· | ···················· | ···················· | ···················· | ···················· | |
| D | 2151 | hCoV-19/USA/CA-CZB-1707/2020\|EPI_ISL_468639\|2020-0 | ···················· | ···················· | ···················· | ···················· | ···················· | ···················· | |
| D | 2152 | hCoV-19/USA/CA-CZB-1709/2020\|EPI_ISL_468640\|2020-0 | ···················· | ···················· | ···················· | ···················· | ···················· | ···················· | |
| D | 2153 | hCoV-19/USA/CA-CZB-1711/2020\|EPI_ISL_468642\|2020-0 | ···················· | ···················· | ···················· | ···················· | ···················· | ···················· | |
| D | 2154 | hCoV-19/USA/CA-CZB-1718/2020\|EPI_ISL_468645\|2020-0 | ···················· | ···················· | ···················· | ···················· | ···················· | ···················· | |
| D | 2155 | hCoV-19/USA/CA-CZB-1720/2020\|EPI_ISL_468646\|2020-0 | ···················· | ···················· | ···················· | ···················· | ···················· | ···················· | |
| D | 2156 | hCoV-19/USA/CA-CZB-1721/2020\|EPI_ISL_468647\|2020-0 | ···················· | ···················· | ···················· | ···················· | ···················· | ···················· | |
| D | 2157 | hCoV-19/USA/CA-CZB-1722/2020\|EPI_ISL_468648\|2020-0 | ···················· | ···················· | ···················· | ···················· | ···················· | ···················· | |
| D | 2158 | hCoV-19/USA/CA-CZB-1726/2020\|EPI_ISL_468649\|2020-0 | ···················· | ···················· | ···················· | ···················· | ···················· | ···················· | |
| D | 2159 | hCoV-19/USA/CA-CZB-1727/2020\|EPI_ISL_468650\|2020-0 | ···················· | ···················· | ···················· | ···················· | ···················· | ···················· | |
| D | 2160 | hCoV-19/USA/CA-CZB-1728/2020\|EPI_ISL_468651\|2020-0 | ···················· | ···················· | ···················· | ···················· | ···················· | ···················· | |
| D | 2161 | hCoV-19/USA/CA-CZB-1729/2020\|EPI_ISL_468652\|2020-0 | ···················· | ···················· | ···················· | ···················· | ···················· | ···················· | |
| D | 2162 | hCoV-19/Croatia/AU-S10new/2020\|EPI_ISL_468656\|2020 | ···················· | ···················· | ···················· | ···················· | ···················· | ···················· | |
| D | 2163 | hCoV-19/England/BRIS-12754F/2020\|EPI_ISL_469792\|20 | ···················· | ·A·················· | ···················· | ···················· | ···················· | ···················· | |
| D | 2164 | hCoV-19/England/BRIS-127837/2020\|EPI_ISL_469793\|20 | ···················· | ···················· | ···················· | ···················· | ···················· | ···················· | |
| D | 2165 | hCoV-19/England/BRIS-127EEA/2020\|EPI_ISL_469798\|20 | ···················· | ···················· | ···················· | ···················· | ···················· | ···················· | |
| D | 2166 | hCoV-19/England/BRIS-12848D/2020\|EPI_ISL_469799\|20 | ···················· | ···················· | ···················· | ···················· | ···················· | ···················· | |
| D | 2167 | hCoV-19/England/CAMB-1B29A7/2020\|EPI_ISL_469903\|20 | ···················· | ···················· | ···················· | ···················· | ···················· | ···················· | |
| D | 2168 | hCoV-19/England/CAMB-1B228E/2020\|EPI_ISL_469906\|20 | ···················· | ···················· | ···················· | ···················· | ···················· | ···················· | |
| D | 2169 | hCoV-19/England/CAMB-1B29C5/2020\|EPI_ISL_469911\|20 | ···················· | ···················· | ···················· | ···················· | ···················· | ···················· | |
| D | 2170 | hCoV-19/England/BRIS-12A3F7/2020\|EPI_ISL_469914\|20 | ···················· | ···················· | ···················· | ···················· | ···················· | ···················· | |
| D | 2171 | hCoV-19/England/NORT-28987E/2020\|EPI_ISL_469915\|20 | ···················· | ···················· | ···················· | ···················· | ···················· | ···················· | |
| D | 2172 | hCoV-19/England/SHEF-10B234D/2020\|EPI_ISL_469918\|2 | ···················· | ···················· | ···················· | ···················· | ···················· | ···················· | |
| D | 2173 | hCoV-19/England/SHEF-10B26BD/2020\|EPI_ISL_469925\|2 | ···················· | ···················· | ···················· | ···················· | ···················· | ···················· | |
| D | 2174 | hCoV-19/England/SHEF-10B22B9/2020\|EPI_ISL_469926\|2 | ···················· | ···················· | ···················· | ···················· | ···················· | ···················· | |
| D | 2175 | hCoV-19/England/SHEF-10B2547/2020\|EPI_ISL_469928\|2 | ···················· | ···················· | ···················· | ···················· | ···················· | ···················· | |
| D | 2176 | hCoV-19/Scotland/GCVR-172202/2020\|EPI_ISL_469930\|2 | ···················· | ···················· | ···················· | ···················· | ···················· | ···················· | |
| D | 2177 | hCoV-19/Scotland/GCVR-172062/2020\|EPI_ISL_469931\|2 | ···················· | ···················· | ···················· | ···················· | ···················· | ···················· | |
| D | 2178 | hCoV-19/Scotland/GCVR-1721F6/2020\|EPI_ISL_469932\|2 | ···················· | ·A·················· | ···················· | ···················· | ···················· | ···················· | |
| D | 2179 | hCoV-19/Scotland/GCVR-172141/2020\|EPI_ISL_469933\|2 | ···················· | ···················· | ···················· | ···················· | ···················· | ···················· | |
| D | 2180 | hCoV-19/Scotland/GCVR-171555/2020\|EPI_ISL_469934\|2 | ···················· | ·A·················· | ···················· | ···················· | ···················· | ···················· | |
| D | 2181 | hCoV-19/Scotland/GCVR-172378/2020\|EPI_ISL_469935\|2 | ···················· | ·A·················· | ···················· | ···················· | ···················· | ···················· | |
| D | 2182 | hCoV-19/Scotland/GCVR-1724B1/2020\|EPI_ISL_469940\|2 | ···················· | ···················· | ···················· | ···················· | ···················· | ···················· | |
| D | 2183 | hCoV-19/Scotland/GCVR-172035/2020\|EPI_ISL_469941\|2 | ···················· | ···················· | ···················· | ···················· | ···················· | ···················· | |
| D | 2184 | hCoV-19/Scotland/GCVR-1721E7/2020\|EPI_ISL_469943\|2 | ···················· | ···················· | ···················· | ···················· | ···················· | ···················· | |
| D | 2185 | hCoV-19/Scotland/GCVR-171F57/2020\|EPI_ISL_469944\|2 | ···················· | ···················· | ···················· | ···················· | ···················· | ···················· | |
| D | 2186 | hCoV-19/Scotland/GCVR-171634/2020\|EPI_ISL_469945\|2 | ···················· | ···················· | ···················· | ···················· | ···················· | ···················· | |
| D | 2187 | hCoV-19/Scotland/GCVR-17224E/2020\|EPI_ISL_469946\|2 | ···················· | ···················· | ···················· | ···················· | ···················· | ···················· | |
| D | 2188 | hCoV-19/Scotland/GCVR-17218D/2020\|EPI_ISL_469947\|2 | ···················· | ···················· | ···················· | ···················· | ···················· | ···················· | |
| D | 2189 | hCoV-19/Scotland/GCVR-172448/2020\|EPI_ISL_469948\|2 | ···················· | ···················· | ···················· | ···················· | ···················· | ···················· | |
| D | 2190 | hCoV-19/Scotland/GCVR-172457/2020\|EPI_ISL_469949\|2 | ········T··········· | ···················· | ···················· | ···················· | ···················· | ···················· | |
| D | 2191 | hCoV-19/Scotland/GCVR-1720BD/2020\|EPI_ISL_469950\|2 | ···················· | ·A·················· | ···················· | ···················· | ···················· | ···················· | |
| D | 2192 | hCoV-19/Scotland/GCVR-172484/2020\|EPI_ISL_469952\|2 | ···················· | ·A·················· | ···················· | ···················· | ···················· | ···················· | |
| D | 2193 | hCoV-19/Scotland/GCVR-172299/2020\|EPI_ISL_469954\|2 | ···················· | ···················· | ···················· | ···················· | ···················· | ···················· | |
| D | 2194 | hCoV-19/Scotland/GCVR-17240C/2020\|EPI_ISL_469956\|2 | ···················· | ···················· | ···················· | ···················· | ···················· | ···················· | |
| D | 2195 | hCoV-19/Scotland/GCVR-1722C6/2020\|EPI_ISL_469957\|2 | ···················· | ···················· | ···················· | ···················· | ···················· | ···················· | |
| D | 2196 | hCoV-19/Scotland/GCVR-171591/2020\|EPI_ISL_469959\|2 | ···················· | ·A·················· | ···················· | ···················· | ···················· | ···················· | |
| D | 2197 | hCoV-19/Scotland/GCVR-172080/2020\|EPI_ISL_469960\|2 | ···················· | ·A·················· | ···················· | ···················· | ···················· | ···················· | |
| D | 2198 | hCoV-19/Scotland/GCVR-172387/2020\|EPI_ISL_469961\|2 | ···················· | ···················· | ···················· | ···················· | ···················· | ···················· | |
| D | 2199 | hCoV-19/Scotland/GCVR-171EF0/2020\|EPI_ISL_469962\|2 | ···················· | ···················A | ···················· | ···················· | ···················· | ···················· | 1 |
| D | 2200 | hCoV-19/Scotland/GCVR-1720DB/2020\|EPI_ISL_469963\|2 | ···················· | ···················· | ···················· | ···················· | ···················· | ···················· | |
| D | 2201 | hCoV-19/Scotland/GCVR-1724C0/2020\|EPI_ISL_469964\|2 | ···················· | ···A················ | ···················· | ···················· | ···················· | ···················· | |
| D | 2202 | hCoV-19/Scotland/GCVR-17227B/2020\|EPI_ISL_469965\|2 | ···················· | ···················· | ···················· | ···················· | ···················· | ···················· | |
| D | 2203 | hCoV-19/Scotland/GCVR-172220/2020\|EPI_ISL_469966\|2 | ···················· | ···················· | ···················· | ···················· | ···················· | ···················· | |
| D | 2204 | hCoV-19/Scotland/GCVR-1723B4/2020\|EPI_ISL_469967\|2 | ···················· | ·A·················· | ···················· | ···················· | ···················· | ···················· | |
| D | 2205 | hCoV-19/Scotland/GCVR-171670/2020\|EPI_ISL_469968\|2 | ···················· | ···················· | ···················· | ···················· | ···················· | ···················· | |
| D | 2206 | hCoV-19/Scotland/GCVR-17226C/2020\|EPI_ISL_469969\|2 | ···················· | ···················· | ···················· | ···················· | ···················· | ···················· | |
| D | 2207 | hCoV-19/Scotland/GCVR-171ED2/2020\|EPI_ISL_469971\|2 | ···················· | ···················· | ···················· | ···················· | ···················· | ···················· | |
| D | 2208 | hCoV-19/Scotland/GCVR-172369/2020\|EPI_ISL_469972\|2 | ···················· | ···················· | ···················· | ···················· | ···················· | ···················· | |
| D | 2209 | hCoV-19/Scotland/GCVR-1721AB/2020\|EPI_ISL_469973\|2 | ···················· | ···················· | ···················· | ···················· | ···················· | ···················· | |
| D | 2210 | hCoV-19/Scotland/GCVR-17242A/2020\|EPI_ISL_469974\|2 | ···················· | ···················· | ···················· | ···················· | ···················· | ···················· | |
| D | 2211 | hCoV-19/Scotland/GCVR-17233C/2020\|EPI_ISL_469976\|2 | ···················· | ·A·················· | ···················· | ···················· | ···················· | ···················· | |
| D | 2212 | hCoV-19/Scotland/GCVR-1720CC/2020\|EPI_ISL_469977\|2 | ···················· | ···················· | ···················· | ···················· | ···················· | ···················· | |
| D | 2213 | hCoV-19/Scotland/GCVR-171F75/2020\|EPI_ISL_469978\|2 | ···················· | ···················A | ···················· | ···················· | ···················· | ···················· | 1 |
| D | 2214 | hCoV-19/Scotland/GCVR-1715FB/2020\|EPI_ISL_469980\|2 | ···················· | ···················· | ···················· | ···················· | ···················· | ···················· | |
| D | 2215 | hCoV-19/Scotland/GCVR-17216F/2020\|EPI_ISL_469981\|2 | ···················· | ···················· | ···················· | ···················· | ···················· | ···················· | |
| D | 2216 | hCoV-19/Scotland/GCVR-172396/2020\|EPI_ISL_469982\|2 | ···················· | ···················· | ···················· | ···················· | ···················· | ···················· | |

```
D   2217 hCoV-19/Scotland/GCVR-1722F3/2020|EPI_ISL_469983|2   .................... .................... .................... .................... .................... ....................
D   2218 hCoV-19/Scotland/GCVR-1715BF/2020|EPI_ISL_469984|2   .................... .................... .................... .................... .................... ....................
D   2219 hCoV-19/Scotland/GCVR-172150/2020|EPI_ISL_469985|2   .................... .................... .................... .................... .................... ....................
D   2220 hCoV-19/Scotland/GCVR-171F0C/2020|EPI_ISL_469986|2   .................... .A.................. .................... .................... .................... ....................
D   2221 hCoV-19/USA/MN-MDH-1077/2020|EPI_ISL_470757|2020-0   .................... .................... .................... .................... .................... ....................
D   2222 hCoV-19/USA/MN-MDH-1078/2020|EPI_ISL_470758|2020-0   .................... .................... .................... .................... .................... ....................
D   2223 hCoV-19/USA/MN-MDH-1080/2020|EPI_ISL_470760|2020-0   .................... .................... .................... .................... .................... ....................
D   2224 hCoV-19/USA/MN-MDH-1081/2020|EPI_ISL_470761|2020-0   .................... .................... .................... .................... .................... ....................
D   2225 hCoV-19/USA/MN-MDH-1082/2020|EPI_ISL_470762|2020-0   .................... .................... .................... .................... .................... ....................
D   2226 hCoV-19/USA/MN-MDH-1083/2020|EPI_ISL_470763|2020-0   .................... .................... .................... .................... .................... ....................
D   2227 hCoV-19/USA/MN-MDH-1084/2020|EPI_ISL_470764|2020-0   .................... .................... .................... .................... .................... ....................
D   2228 hCoV-19/USA/MN-MDH-1085/2020|EPI_ISL_470765|2020-0   .................... .................... .................... .................... .................... ....................
D   2229 hCoV-19/USA/MN-MDH-1086/2020|EPI_ISL_470766|2020-0   .................... .................... .................... .................... .................... ....................
D   2230 hCoV-19/USA/MN-MDH-1087/2020|EPI_ISL_470767|2020-0   .................... .................... .................... .................... .................... ....................
D   2231 hCoV-19/USA/MN-MDH-1088/2020|EPI_ISL_470768|2020-0   .................... .................... .................... .................... .................... ....................
D   2232 hCoV-19/USA/MN-MDH-1089/2020|EPI_ISL_470769|2020-0   .................... .................... .................... .................... .................... ....................
D   2233 hCoV-19/Russia/Moscow_PMVL-7/2020|EPI_ISL_470900|2   .................... .................... .................... .................... .................... ....................
D   2234 hCoV-19/Russia/Moscow_PMVL-8/2020|EPI_ISL_470901|2   .................... .................... .................... .................... .................... ....................
D   2235 hCoV-19/Russia/Moscow_PMVL-9/2020|EPI_ISL_470902|2   .................... .................... .................... .................... .................... ....................
D   2236 hCoV-19/Russia/Moscow_PMVL-10/2020|EPI_ISL_470903|   .................... .................... .................... .................... .................... ....................
D   2237 hCoV-19/Russia/Moscow_PMVL-11/2020|EPI_ISL_470904|   .................... .................... .................... .................... .................... ....................
D   2238 hCoV-19/Wales/PHWC-162803/2020|EPI_ISL_473125|2020   .................... .................... .................... .................... .................... ....................
D   2239 hCoV-19/Wales/PHWC-162812/2020|EPI_ISL_473126|2020   .................... .................... .................... .................... .................... ....................
D   2240 hCoV-19/Wales/PHWC-162830/2020|EPI_ISL_473127|2020   .................... .................... .................... .................... .................... ....................
D   2241 hCoV-19/Wales/PHWC-16288B/2020|EPI_ISL_473132|2020   .................... .................... .................... .................... .................... ....................
D   2242 hCoV-19/Wales/PHWC-1628A9/2020|EPI_ISL_473134|2020   .................... .................... .................... .................... .................... ....................
D   2243 hCoV-19/Wales/PHWC-1628F4/2020|EPI_ISL_473139|2020   .................... .................... .................... .................... .................... ....................
D   2244 hCoV-19/Wales/PHWC-16291F/2020|EPI_ISL_473141|2020   .................... .................... .................... .................... .................... ....................
D   2245 hCoV-19/Wales/PHWC-162988/2020|EPI_ISL_473146|2020   .................... .................... .................... .................... .................... ....................
D   2246 hCoV-19/Wales/PHWC-1629C4/2020|EPI_ISL_473147|2020   .................... .................... .................... .................... .................... ....................
D   2247 hCoV-19/Wales/PHWC-1629E2/2020|EPI_ISL_473149|2020   .................... .................... .................... .................... .................... ....................
D   2248 hCoV-19/Wales/PHWC-162A67/2020|EPI_ISL_473155|2020   .................... .................... .................... .................... .................... ....................
D   2249 hCoV-19/Wales/PHWC-162AB2/2020|EPI_ISL_473158|2020   .................... .................... .................... .................... .................... ....................
D   2250 hCoV-19/Wales/PHWC-162AFE/2020|EPI_ISL_473162|2020   .................... .................... .................... .................... .................... ....................
D   2251 hCoV-19/Wales/PHWC-162B0A/2020|EPI_ISL_473163|2020   .................... .................... .................... .................... .................... ....................
D   2252 hCoV-19/Wales/PHWC-162B37/2020|EPI_ISL_473166|2020   .................... .................... .................... .................... .................... ....................
D   2253 hCoV-19/Wales/PHWC-162B55/2020|EPI_ISL_473168|2020   .................... .................... .................... .................... .................... ....................
D   2254 hCoV-19/Wales/PHWC-162B73/2020|EPI_ISL_473170|2020   .................... .................... .................... .................... .................... ....................
D   2255 hCoV-19/Wales/PHWC-162BA0/2020|EPI_ISL_473173|2020   .................... .................... .................... .................... .................... ....................
D   2256 hCoV-19/Wales/PHWC-162C07/2020|EPI_ISL_473178|2020   .................... .................... .................... .................... .................... ....................
D   2257 hCoV-19/Wales/PHWC-162C16/2020|EPI_ISL_473179|2020   .................... .................... .................... .................... .................... ....................
D   2258 hCoV-19/Wales/PHWC-162C70/2020|EPI_ISL_473183|2020   .................... .................... .................... .................... .................... ....................
D   2259 hCoV-19/Wales/PHWC-162CCB/2020|EPI_ISL_473186|2020   .................... .................... .................... .................... .................... ....................
D   2260 hCoV-19/Wales/PHWC-162CE9/2020|EPI_ISL_473188|2020   .................... .................... .................... .................... .................... ....................
D   2261 hCoV-19/Wales/PHWC-162CF8/2020|EPI_ISL_473189|2020   .................... ...A................ .................... .................... .................... ....................
D   2262 hCoV-19/Wales/PHWC-162D40/2020|EPI_ISL_473194|2020   .................... .................... .................... .................... .................... ....................
D   2263 hCoV-19/Wales/PHWC-162D8C/2020|EPI_ISL_473198|2020   .................... .................... .................... .................... .................... ....................
D   2264 hCoV-19/Wales/PHWC-162D9B/2020|EPI_ISL_473199|2020   .................... .................... .................... .................... .................... ....................
D   2265 hCoV-19/Wales/PHWC-3570D/2020|EPI_ISL_474464|2020-   .................... .................... .................... .................... .................... ....................
D   2266 hCoV-19/Wales/PHWC-3572B/2020|EPI_ISL_474466|2020-   .................... .................... .................... .................... .................... ....................
D   2267 hCoV-19/Wales/PHWC-3573A/2020|EPI_ISL_474467|2020-   .................... .................... .................... .................... .................... ....................
D   2268 hCoV-19/Wales/PHWC-35749/2020|EPI_ISL_474468|2020-   .................... .................... .................... .................... .................... ....................
D   2269 hCoV-19/Wales/PHWC-35776/2020|EPI_ISL_474471|2020-   .................... .................... .................... .................... .................... ....................
D   2270 hCoV-19/Wales/PHWC-357D0/2020|EPI_ISL_474477|2020-   .................... .................... .................... .................... .................... ....................
D   2271 hCoV-19/Wales/PHWC-357EF/2020|EPI_ISL_474478|2020-   .................... .................... .................... .................... .................... ....................
D   2272 hCoV-19/Wales/PHWC-357FE/2020|EPI_ISL_474479|2020-   .................... .................... .................... .................... .................... ....................
D   2273 hCoV-19/Wales/PHWC-3580A/2020|EPI_ISL_474480|2020-   .................... .................... .................... .................... .................... ....................
D   2274 hCoV-19/Wales/PHWC-35864/2020|EPI_ISL_474486|2020-   .................... .................... .................... .................... .................... ....................
D   2275 hCoV-19/Wales/PHWC-358A0/2020|EPI_ISL_474490|2020-   .................... .................... .................... .................... .................... ....................
D   2276 hCoV-19/Wales/PHWC-358EC/2020|EPI_ISL_474494|2020-   .................... .................... .................... .................... .................... ....................
D   2277 hCoV-19/Wales/PHWC-35916/2020|EPI_ISL_474497|2020-   .................... .................... .................... .................... .................... ....................
D   2278 hCoV-19/Wales/PHWC-35925/2020|EPI_ISL_474498|2020-   .................... .................... .................... .................... .................... ....................
D   2279 hCoV-19/USA/UT-04105/2020|EPI_ISL_475743|2020-05-1   .................... .................... .................... .................... .................... ....................
D   2280 hCoV-19/Egypt/MASRI-2/2020|EPI_ISL_475745|2020-05     .................... .....A.............. .................... .................... .................... ....................
D   2281 hCoV-19/Egypt/MASRI-3/2020|EPI_ISL_475746|2020-05     .................... .................... .................... .................... .................... ....................
D   2282 hCoV-19/Egypt/MASRI-6/2020|EPI_ISL_475747|2020-05     .................... .................... .................... .................... .................... ....................
D   2283 hCoV-19/Egypt/MASRI-13/2020|EPI_ISL_475748|2020-05   .................... .....A.............. .................... .................... .................... ....................
D   2284 hCoV-19/Egypt/MASRI-14/2020|EPI_ISL_475749|2020-05   .................... .................... .................... .................... .................... ....................
D   2285 hCoV-19/Egypt/MASRI-9/2020|EPI_ISL_475751|2020-05     .................... ........A........... .................... .................... .................... ....................
D   2286 hCoV-19/Austria/CeMM0313/2020|EPI_ISL_475785|2020-   .................... .................... .................... .................... .................... ....................
D   2287 hCoV-19/Austria/CeMM0314/2020|EPI_ISL_475786|2020-   .................... .................... .................... .................... .................... ....................
D   2288 hCoV-19/Austria/CeMM0315/2020|EPI_ISL_475787|2020-   .................... .................... .................... .................... .................... ....................
D   2289 hCoV-19/Austria/CeMM0319/2020|EPI_ISL_475790|2020-   .................... .................... .................... .................... .................... ....................
D   2290 hCoV-19/Austria/CeMM0320/2020|EPI_ISL_475791|2020-   .................... .................... .................... .................... .................... ....................
D   2291 hCoV-19/Austria/CeMM0321/2020|EPI_ISL_475792|2020-   .................... .................... .................... .................... .................... ....................
D   2292 hCoV-19/Austria/CeMM0323/2020|EPI_ISL_475793|2020-   .................... .................... .................... .................... .................... ....................
D   2293 hCoV-19/Austria/CeMM0324/2020|EPI_ISL_475794|2020-   .................... .................... .................... .................... .................... ....................
D   2294 hCoV-19/Austria/CeMM0326/2020|EPI_ISL_475795|2020-   .................... .................... .................... .................... .................... ....................
```

| | | | | | | | | |
|---|---|---|---|---|---|---|---|---|
| D | 2295 | hCoV-19/Austria/CeMM0330/2020\|EPI_ISL_475797\|2020- | ···················· | ···················· | ···················· | ···················· | ···················· | ···················· |
| D | 2296 | hCoV-19/Austria/CeMM0331/2020\|EPI_ISL_475798\|2020- | ···················· | ···················· | ···················· | ···················· | ···················· | ···················· |
| D | 2297 | hCoV-19/Austria/CeMM0333/2020\|EPI_ISL_475799\|2020- | ···················· | ···················· | ···················· | ···················· | ···················· | ···················· |
| D | 2298 | hCoV-19/England/OXON-B1EE6/2020\|EPI_ISL_479100\|202 | ···················· | ···················· | ···················· | ···················· | ···················· | ···················· |
| D | 2299 | hCoV-19/England/OXON-B1EF5/2020\|EPI_ISL_479101\|202 | ···················· | ···················· | ···················· | ···················· | ···················· | ···················· |
| D | 2300 | hCoV-19/England/OXON-B1F2F/2020\|EPI_ISL_479102\|202 | ···················· | ···················· | ···················· | ···················· | ···················· | ···················· |
| D | 2301 | hCoV-19/England/OXON-B1F98/2020\|EPI_ISL_479103\|202 | ···················· | ···················· | ···················· | ···················· | ···················· | ···················· |
| D | 2302 | hCoV-19/England/OXON-B1FC5/2020\|EPI_ISL_479104\|202 | ···················· | ···················· | ···················· | ···················· | ···················· | ···················· |
| D | 2303 | hCoV-19/England/OXON-B1FF2/2020\|EPI_ISL_479105\|202 | ···················· | ···················· | ···················· | ···················· | ···················· | ···················· |
| D | 2304 | hCoV-19/England/OXON-B201C/2020\|EPI_ISL_479106\|202 | ···················· | ···················· | ···················· | ···················· | ···················· | ···················· |
| D | 2305 | hCoV-19/Singapore/282/2020\|EPI_ISL_462400\|2020-04- | ···················· | ···················· | ···················· | ···················· | ···················· | ···················· |
| D | 2306 | hCoV-19/Singapore/283/2020\|EPI_ISL_462401\|2020-03- | ···················· | ···················· | ···················· | ···················· | ···················· | ···················· |
| D | 2307 | hCoV-19/Singapore/284/2020\|EPI_ISL_462402\|2020-04- | ···················· | ···················· | ···················· | ···················· | ···················· | ···················· |
| D | 2308 | hCoV-19/Singapore/285/2020\|EPI_ISL_462403\|2020-04- | ···················· | ···················· | ···················· | ···················· | ···················· | ···················· |
| D | 2309 | hCoV-19/Singapore/286/2020\|EPI_ISL_462404\|2020-03- | ···················· | ···················· | ···················· | ···················· | ···················· | ···················· |
| D | 2310 | hCoV-19/Singapore/287/2020\|EPI_ISL_462405\|2020-03- | ···················· | ···················· | ···················· | ···················· | ···················· | ···················· |
| D | 2311 | hCoV-19/Singapore/288/2020\|EPI_ISL_462406\|2020-04- | ···················· | ···················· | ···················· | ···················· | ···················· | ···················· |
| D | 2312 | hCoV-19/Singapore/289/2020\|EPI_ISL_462407\|2020-04- | ···················· | ···················· | ···················· | ···················· | ···················· | ···················· |
| D | 2313 | hCoV-19/Singapore/290/2020\|EPI_ISL_462408\|2020-04- | ···················· | ···················· | ···················· | ···················· | ···················· | ···················· |
| D | 2314 | hCoV-19/Singapore/291/2020\|EPI_ISL_462409\|2020-03- | ···················· | ···················· | ···················· | ···················· | ···················· | ···················· |
| D | 2315 | hCoV-19/Singapore/292/2020\|EPI_ISL_462410\|2020-03- | ···················· | ···················· | ···················· | ···················· | ···················· | ···················· |
| D | 2316 | hCoV-19/Singapore/293/2020\|EPI_ISL_462411\|2020-04- | ···················· | ···················· | ···················· | ···················· | ···················· | ···················· |
| D | 2317 | hCoV-19/Singapore/294/2020\|EPI_ISL_462412\|2020-05- | ···················· | ···················· | ···················· | ···················· | ···················· | ···················· |
| D | 2318 | hCoV-19/Singapore/295/2020\|EPI_ISL_462413\|2020-04- | ···················· | ···················· | ···················· | ···················· | ···················· | ···················· |
| D | 2319 | hCoV-19/Singapore/296/2020\|EPI_ISL_462414\|2020-04- | ···················· | ···················· | ···················· | ···················· | ···················· | ···················· |
| D | 2320 | hCoV-19/Singapore/297/2020\|EPI_ISL_462415\|2020-03- | ···················· | ···················· | ···················· | ···················· | ···················· | ···················· |
| D | 2321 | hCoV-19/Singapore/298/2020\|EPI_ISL_462416\|2020-03- | ···················· | ···················· | ···················· | ···················· | ···················· | ···················· |
| D | 2322 | hCoV-19/Singapore/299/2020\|EPI_ISL_462417\|2020-04- | ···················· | ···················· | ···················· | ···················· | ···················· | ···················· |
| D | 2323 | hCoV-19/Singapore/300/2020\|EPI_ISL_462418\|2020-04- | ···················· | ···················· | ···················· | ···················· | ···················· | ···················· |
| D | 2324 | hCoV-19/Singapore/301/2020\|EPI_ISL_462419\|2020-04- | ···················· | ···················· | ···················· | ···················· | ···················· | ···················· |
| D | 2325 | hCoV-19/Singapore/302/2020\|EPI_ISL_462420\|2020-03- | ···················· | ···················· | ···················· | ···················· | ···················· | ···················· |
| D | 2326 | hCoV-19/Singapore/303/2020\|EPI_ISL_462421\|2020-04- | ···················· | ···················· | ···················· | ···················· | ···················· | ···················· |
| D | 2327 | hCoV-19/Singapore/304/2020\|EPI_ISL_462422\|2020-03- | ···················· | ···················· | ···················· | ···················· | ···················· | ···················· |
| D | 2328 | hCoV-19/Singapore/305/2020\|EPI_ISL_462423\|2020-04- | ···················· | ···················· | ···················· | ···················· | ···················· | ···················· |
| D | 2329 | hCoV-19/Singapore/306/2020\|EPI_ISL_462424\|2020-04- | ···················· | ···················· | ···················· | ···················· | ···················· | ···················· |
| D | 2330 | hCoV-19/Singapore/307/2020\|EPI_ISL_462425\|2020-04- | ···················· | ···················· | ···················· | ···················· | ···················· | ···················· |
| D | 2331 | hCoV-19/Singapore/308/2020\|EPI_ISL_462426\|2020-04- | ···················· | ···················· | ···················· | ···················· | ···················· | ···················· |
| D | 2332 | hCoV-19/Singapore/309/2020\|EPI_ISL_462427\|2020-05- | ···················· | ···················· | ···················· | ···················· | ···················· | ···················· |
| D | 2333 | hCoV-19/Singapore/310/2020\|EPI_ISL_462428\|2020-04- | ···················· | ···················· | ···················· | ···················· | ···················· | ···················· |
| D | 2334 | hCoV-19/Singapore/311/2020\|EPI_ISL_462429\|2020-03- | ···················· | ···················· | ···················· | ···················· | ···················· | ···················· |
| D | 2335 | hCoV-19/Singapore/312/2020\|EPI_ISL_462430\|2020-04- | ···················· | ···················· | ···················· | ···················· | ···················· | ···················· |
| D | 2336 | hCoV-19/Bosnia and Herzegovina/ChVir7377/2020\|EPI_ | ···················· | ···················· | ···················· | ···················· | ···················· | ···················· |
| D | 2337 | hCoV-19/Bosnia and Herzegovina/ChVir7379/2020\|EPI_ | ···················· | ···················· | ···················· | ···················· | ···················· | ···················· |
| D | 2338 | hCoV-19/Spain/MD-ISCIII-201276/2020\|EPI_ISL_462478 | ···················· | ···················· | ···················· | ···················· | ···················· | ···················· |
| D | 2339 | hCoV-19/Spain/CT-ISCIII-202052/2020\|EPI_ISL_462479 | ···················· | ···················· | ···················· | ···················· | ···················· | ···················· |
| D | 2340 | hCoV-19/Poland/IHG_PAS_3_57/2020\|EPI_ISL_462480\|20 | ···················· | ···················· | ···················· | ···················· | ···················· | ···················· |
| D | 2341 | hCoV-19/USA/WA-S1210/2020\|EPI_ISL_463700\|2020-05-0 | ···················· | ···················· | ···················· | ···················· | ···················· | ···················· |
| D | 2342 | hCoV-19/USA/WA-S1211/2020\|EPI_ISL_463701\|2020-05-0 | ···················· | ···················· | ···················· | ···················· | ···················· | ···················· |
| D | 2343 | hCoV-19/USA/WA-S1212/2020\|EPI_ISL_463702\|2020-05-0 | ···················· | ···················· | ···················· | ···················· | ···················· | ···················· |
| D | 2344 | hCoV-19/USA/WA-S1213/2020\|EPI_ISL_463703\|2020-05-0 | ···················· | ···················· | ···················· | ···················· | ···················· | ···················· |
| D | 2345 | hCoV-19/USA/WA-S1214/2020\|EPI_ISL_463704\|2020-05-0 | ···················· | ···················· | ···················· | ···················· | ···················· | ···················· |
| D | 2346 | hCoV-19/USA/WA-S1215/2020\|EPI_ISL_463705\|2020-05-0 | ···················· | ···················· | ···················· | ···················· | ···················· | ···················· |
| D | 2347 | hCoV-19/United Arab Emirates/P1/2020\|EPI_ISL_46374 | ···················· | ···················· | ···················· | ···················· | ·········T·········· | ···················· |
| D | 2348 | hCoV-19/Cyprus/001/2020\|EPI_ISL_463741\|2020-03-11 | ···················· | ···················· | ···················· | ···················· | ···················· | ···················· |
| D | 2349 | hCoV-19/Cyprus/002/2020\|EPI_ISL_463742\|2020-03-22 | ···················· | ···················· | ···················· | ···················· | ···················· | ···················· |
| D | 2350 | hCoV-19/Cyprus/003/2020\|EPI_ISL_463743\|2020-03-25 | ···················· | ···················· | ···················· | ···················· | ···················· | ···················· |
| D | 2351 | hCoV-19/Cyprus/004/2020\|EPI_ISL_463744\|2020-03-23 | ···················· | ···················· | ···················· | ···················· | ···················· | ···················· |
| D | 2352 | hCoV-19/Cyprus/005/2020\|EPI_ISL_463745\|2020-04-01 | ···················· | ···················· | ···················· | ···················· | ···················· | ···················· |
| D | 2353 | hCoV-19/Cyprus/006/2020\|EPI_ISL_463746\|2020-04-11 | ···················· | ···················· | ···················· | ···················· | ···················· | ···················· |
| D | 2354 | hCoV-19/Cyprus/007/2020\|EPI_ISL_463747\|2020-04-17 | ···················· | ···················· | ···················· | ···················· | ···················· | ···················· |
| D | 2355 | hCoV-19/Cyprus/008/2020\|EPI_ISL_463748\|2020-04-27 | ···················· | ···················· | ···················· | ···················· | ···················· | ···················· |
| D | 2357 | hCoV-19/Spain/MD-IBV-003662/2020\|EPI_ISL_467206\|20 | ···················· | ···················· | ···················· | ···················· | ···················· | ···················· |
| D | 2358 | hCoV-19/Spain/MD-IBV-003697/2020\|EPI_ISL_467207\|20 | ···················· | ···················· | ···················· | ···················· | ···················· | ···················· |
| D | 2360 | hCoV-19/Spain/MD-IBV-003668/2020\|EPI_ISL_467209\|20 | ···················· | ···················· | ···················· | ···················· | ···················· | ···················· |
| D | 2363 | hCoV-19/Spain/MD-IBV-003653/2020\|EPI_ISL_467213\|20 | ···················· | ···················· | ···················· | ···················· | ···················· | ···················· |
| D | 2365 | hCoV-19/Spain/MD-IBV-003665/2020\|EPI_ISL_467215\|20 | ···················· | ···················· | ···················· | ···················· | ···················· | ···················· |
| D | 2366 | hCoV-19/Spain/MD-IBV-003709/2020\|EPI_ISL_467216\|20 | ···················· | ···················· | ···················· | ···················· | ···················· | ···················· |
| D | 2369 | hCoV-19/Spain/MD-IBV-003681/2020\|EPI_ISL_467219\|20 | ···················· | ···················· | ···················· | ···················· | ···················· | ···················· |
| D | 2372 | hCoV-19/Spain/MD-IBV-003660/2020\|EPI_ISL_467222\|20 | ···················· | ···················· | ···················· | ···················· | ···················· | ···················· |
| D | 2374 | hCoV-19/Spain/MD-IBV-003706/2020\|EPI_ISL_467224\|20 | ···················· | ···················· | ···················· | ···················· | ···················· | ···················· |
| D | 2377 | hCoV-19/Spain/MD-IBV-003707/2020\|EPI_ISL_467227\|20 | ···················· | ···················· | ···················· | ···················· | ···················· | ···················· |
| D | 2379 | hCoV-19/Spain/MD-IBV-003694/2020\|EPI_ISL_467229\|20 | ···················· | ···················· | ···················· | ···················· | ···················· | ···················· |
| D | 2380 | hCoV-19/Spain/MD-IBV-003677/2020\|EPI_ISL_467230\|20 | ···················· | ···················· | ···················· | ···················· | ···················· | ···················· |
| D | 2384 | hCoV-19/Spain/MD-IBV-003723/2020\|EPI_ISL_467234\|20 | ···················· | ···················· | ···················· | ···················· | ···················· | ···················· |
| D | 2385 | hCoV-19/Spain/MD-IBV-003702/2020\|EPI_ISL_467235\|20 | ···················· | ···················· | ···················· | ···················· | ···················· | ···················· |
| D | 2389 | hCoV-19/Spain/MD-IBV-003657/2020\|EPI_ISL_467239\|20 | ···················· | ···················· | ···················· | ···················· | ···················· | ···················· |
| D | 2391 | hCoV-19/Spain/MD-IBV-003717/2020\|EPI_ISL_467241\|20 | ···················· | ···················· | ···················· | ···················· | ···················· | ···················· |
| D | 2395 | hCoV-19/Spain/MD-IBV-003661/2020\|EPI_ISL_467245\|20 | ···················· | ···················· | ···················· | ···················· | ···················· | ···················· |

| | | | | | | | |
|---|---|---|---|---|---|---|---|
| D | 2396 hCoV-19/Spain/MD-IBV-003654/2020\|EPI_ISL_467247\|20 | .................... | .................... | ..................... | .................... | .................... | .................... |
| D | 2407 hCoV-19/Spain/GA-IBV-002733/2020\|EPI_ISL_467289\|20 | .................... | .................... | ..................... | .................... | .................... | .................... |
| D | 2413 hCoV-19/Spain/GA-IBV-002723/2020\|EPI_ISL_467295\|20 | .................... | .................... | ..................... | .................... | .................... | .................... |
| D | 2416 hCoV-19/USA/CA-CZB-1607/2020\|EPI_ISL_468536\|2020-0 | .................... | .................... | ..................... | .................... | .................... | .................... |
| D | 2417 hCoV-19/USA/CA-CZB-1608/2020\|EPI_ISL_468537\|2020-0 | .................... | .................... | ..................... | .................... | .................... | .................... |
| D | 2418 hCoV-19/USA/CA-CZB-1609/2020\|EPI_ISL_468538\|2020-0 | .................... | .................... | ..................... | .................... | .................... | .................... |
| D | 2419 hCoV-19/USA/CA-CZB-1610/2020\|EPI_ISL_468539\|2020-0 | .................... | .................... | ..................... | .................... | .................... | .................... |
| D | 2420 hCoV-19/USA/CA-CZB-1611/2020\|EPI_ISL_468540\|2020-0 | .................... | .................... | ..................... | .................... | .................... | .................... |
| D | 2421 hCoV-19/USA/CA-CZB-1612/2020\|EPI_ISL_468541\|2020-0 | .................... | .................... | ..................... | .................... | .................... | .................... |
| D | 2422 hCoV-19/USA/CA-CZB-1613/2020\|EPI_ISL_468542\|2020-0 | .................... | .................... | ..................... | .................... | .................... | .................... |
| D | 2423 hCoV-19/USA/CA-CZB-1614/2020\|EPI_ISL_468543\|2020-0 | .................... | .................... | ..................... | .................... | .................... | .................... |
| D | 2424 hCoV-19/USA/CA-CZB-1615/2020\|EPI_ISL_468544\|2020-0 | .................... | .................... | ..................... | .................... | .................... | .................... |
| D | 2425 hCoV-19/USA/CA-CZB-1616/2020\|EPI_ISL_468545\|2020-0 | .................... | .................... | ..................... | .................... | .................... | .................... |
| D | 2426 hCoV-19/USA/CA-CZB-1617/2020\|EPI_ISL_468546\|2020-0 | .................... | .................... | ..................... | .................... | .................... | .................... |
| D | 2427 hCoV-19/USA/CA-CZB-1618/2020\|EPI_ISL_468547\|2020-0 | .................... | .................... | ..................... | .................... | .................... | .................... |
| D | 2428 hCoV-19/USA/CA-CZB-1619/2020\|EPI_ISL_468548\|2020-0 | .................... | .................... | ..................... | .................... | .................... | .................... |
| D | 2429 hCoV-19/USA/CA-CZB-1620/2020\|EPI_ISL_468549\|2020-0 | .................... | .................... | ..................... | .................... | .................... | .................... |
| D | 2430 hCoV-19/USA/CA-CZB-1622/2020\|EPI_ISL_468551\|2020-0 | .................... | .................... | ..................... | .................... | .................... | .................... |
| D | 2431 hCoV-19/USA/CA-CZB-1623/2020\|EPI_ISL_468552\|2020-0 | .................... | .................... | ..................... | .................... | .................... | .................... |
| D | 2432 hCoV-19/USA/CA-CZB-1624/2020\|EPI_ISL_468553\|2020-0 | .................... | .................... | ..................... | .................... | .................... | .................... |
| D | 2433 hCoV-19/USA/CA-CZB-1625/2020\|EPI_ISL_468554\|2020-0 | .................... | .................... | ..................... | .................... | .................... | .................... |
| D | 2434 hCoV-19/USA/CA-CZB-1628/2020\|EPI_ISL_468557\|2020-0 | .................... | .A.................. | ..................... | .................... | .................... | .................... |
| D | 2435 hCoV-19/USA/CA-CZB-1629/2020\|EPI_ISL_468558\|2020-0 | .................... | .................... | ..................... | .................... | .................... | .................... |
| D | 2436 hCoV-19/USA/CA-QDX-101/2020\|EPI_ISL_468560\|2020-03 | .................... | .................... | ..................... | .................... | .................... | .................... |
| D | 2437 hCoV-19/USA/CA-QDX-103/2020\|EPI_ISL_468561\|2020-03 | .................... | .................... | ..................... | .................... | .................... | .................... |
| D | 2438 hCoV-19/USA/CA-QDX-110/2020\|EPI_ISL_468562\|2020-03 | .................... | .................... | ..................... | .................... | .................... | .................... |
| D | 2439 hCoV-19/USA/CA-QDX-111/2020\|EPI_ISL_468563\|2020-03 | .................... | .................... | ..................... | .................... | .................... | .................... |
| D | 2440 hCoV-19/USA/CA-QDX-83/2020\|EPI_ISL_468564\|2020-03- | .................... | .................... | ..................... | .................... | .................... | .................... |
| D | 2441 hCoV-19/USA/CA-QDX-84/2020\|EPI_ISL_468565\|2020-03- | .................... | .................... | ..................... | .................... | .................... | .................... |
| D | 2442 hCoV-19/USA/CA-QDX-86/2020\|EPI_ISL_468566\|2020-03- | .................... | .................... | ..................... | .................... | .................... | .................... |
| D | 2443 hCoV-19/USA/CA-QDX-92/2020\|EPI_ISL_468567\|2020-03- | .................... | .................... | ..................... | .................... | .................... | .................... |
| D | 2444 hCoV-19/USA/CA-QDX-97/2020\|EPI_ISL_468568\|2020-03- | .................... | .................... | ..................... | .................... | .................... | .................... |
| D | 2445 hCoV-19/USA/CA-QDX-98/2020\|EPI_ISL_468569\|2020-03- | .................... | .................... | ..................... | .................... | .................... | .................... |
| D | 2446 hCoV-19/USA/CA-QDX-99/2020\|EPI_ISL_468570\|2020-03- | .................... | .................... | ..................... | .................... | .................... | .................... |
| D | 2447 hCoV-19/USA/CO-QDX-107/2020\|EPI_ISL_468571\|2020-03 | .................... | .................... | ..................... | .................... | .................... | .................... |
| D | 2448 hCoV-19/USA/CO-QDX-89/2020\|EPI_ISL_468572\|2020-03- | .................... | .................... | ..................... | .................... | .................... | .................... |
| D | 2449 hCoV-19/USA/HI-QDX-163/2020\|EPI_ISL_468573\|2020-03 | .................... | .................... | ..................... | .................... | .................... | .................... |
| D | 2450 hCoV-19/USA/HI-QDX-164/2020\|EPI_ISL_468574\|2020-03 | .................... | .................... | ..................... | .................... | .................... | .................... |
| D | 2451 hCoV-19/USA/IN-QDX-85/2020\|EPI_ISL_468575\|2020-03- | .................... | .................... | ..................... | .................... | .................... | .................... |
| D | 2452 hCoV-19/USA/NY-QDX-100/2020\|EPI_ISL_468576\|2020-03 | .................... | .................... | ..................... | .................... | .................... | .................... |
| D | 2453 hCoV-19/USA/NY-QDX-102/2020\|EPI_ISL_468577\|2020-03 | .................... | .................... | ..................... | .................... | .................... | .................... |
| D | 2454 hCoV-19/USA/NY-QDX-105/2020\|EPI_ISL_468578\|2020-03 | .................... | .................... | ..................... | .................... | .................... | .................... |
| D | 2455 hCoV-19/USA/NY-QDX-106/2020\|EPI_ISL_468579\|2020-03 | .................... | .................... | ..................... | .................... | .................... | .................... |
| D | 2456 hCoV-19/Northern Ireland/NIRE-101763/2020\|EPI_ISL_ | .................... | .................... | ..................... | .................... | .................... | .................... |
| D | 2457 hCoV-19/Northern Ireland/NIRE-102726/2020\|EPI_ISL_ | .................... | .................... | ..................... | .................... | .................... | .................... |
| D | 2458 hCoV-19/Northern Ireland/NIRE-1025FF/2020\|EPI_ISL_ | .................... | .................... | ..................... | .................... | .................... | .................... |
| D | 2459 hCoV-19/Northern Ireland/NIRE-1015A5/2020\|EPI_ISL_ | .................... | .................... | ..................... | .................... | .................... | .................... |
| D | 2460 hCoV-19/Northern Ireland/NIRE-10145D/2020\|EPI_ISL_ | .................... | .................... | ..................... | .................... | .................... | .................... |
| D | 2461 hCoV-19/Northern Ireland/NIRE-FF2D1/2020\|EPI_ISL_4 | .................... | .................... | ..................... | .................... | .................... | .................... |
| D | 2462 hCoV-19/Northern Ireland/NIRE-10252C/2020\|EPI_ISL_ | .................... | .................... | ..................... | .................... | .................... | .................... |
| D | 2463 hCoV-19/Northern Ireland/NIRE-FF3CF/2020\|EPI_ISL_4 | .................... | .................... | ..................... | .................... | .................... | .................... |
| D | 2464 hCoV-19/Northern Ireland/NIRE-FF471/2020\|EPI_ISL_4 | .................... | .................... | ..................... | .................... | .................... | .................... |
| D | 2465 hCoV-19/Northern Ireland/NIRE-1010D1/2020\|EPI_ISL_ | .................... | .A.................. | ..................... | .................... | .................... | .................... |
| D | 2466 hCoV-19/Northern Ireland/NIRE-100DDB/2020\|EPI_ISL_ | .................... | .................... | ..................... | .................... | .................... | .................... |
| D | 2467 hCoV-19/Northern Ireland/NIRE-101569/2020\|EPI_ISL_ | .................... | .................... | ..................... | .................... | .................... | .................... |
| D | 2468 hCoV-19/Northern Ireland/NIRE-100C74/2020\|EPI_ISL_ | .................... | .................... | ..................... | .................... | .................... | .................... |
| D | 2469 hCoV-19/Northern Ireland/NIRE-FF7E1/2020\|EPI_ISL_4 | .................... | .................... | ..................... | .................... | .................... | .................... |
| D | 2470 hCoV-19/England/CAMB-1B1F1F/2020\|EPI_ISL_469896\|20 | .................... | .................... | ..................... | .................... | .................... | .................... |
| D | 2471 hCoV-19/England/CAMB-1B1EF4/2020\|EPI_ISL_469899\|20 | .................... | .................... | ..................... | .................... | .................... | .................... |
| D | 2472 hCoV-19/Australia/WA34/2020\|EPI_ISL_470851\|2020-02 | .................... | .................... | ..................... | .................... | .................... | .................... |
| D | 2473 hCoV-19/Australia/WA36/2020\|EPI_ISL_470853\|2020-03 | .................... | .................... | ..................... | .................... | .................... | .................... |
| D | 2474 hCoV-19/Australia/WA37/2020\|EPI_ISL_470854\|2020-04 | .................... | .................... | ..................... | .................... | .................... | .................... |
| D | 2475 hCoV-19/Australia/WA38/2020\|EPI_ISL_470855\|2020-04 | .................... | .................... | ..................... | .................... | .................... | .................... |
| D | 2476 hCoV-19/Australia/WA39/2020\|EPI_ISL_470856\|2020-03 | .................... | .................... | ..................... | .................... | .................... | .................... |
| D | 2477 hCoV-19/Australia/WA41/2020\|EPI_ISL_470858\|2020-03 | .................... | .................... | ..................... | .................... | .................... | .................... |
| D | 2478 hCoV-19/Australia/WA44/2020\|EPI_ISL_470861\|2020-04 | .................... | .................... | ..................... | .................... | .................... | .................... |
| D | 2479 hCoV-19/Australia/WA45/2020\|EPI_ISL_470862\|2020-04 | .................... | .................... | ..................... | .................... | .................... | .................... |
| D | 2480 hCoV-19/Australia/WA46/2020\|EPI_ISL_470863\|2020-03 | .................... | .................... | ..................... | .................... | .................... | .................... |
| D | 2481 hCoV-19/Australia/WA47/2020\|EPI_ISL_470864\|2020-03 | .................... | .................... | ..................... | .................... | .................... | .................... |
| D | 2482 hCoV-19/Russia/Moscow_PMVL-2/2020\|EPI_ISL_470896\|2 | .................... | .................... | ..................... | .................... | .................... | .................... |
| D | 2483 hCoV-19/Russia/Moscow_PMVL-3/2020\|EPI_ISL_470897\|2 | .................... | .................... | ..................... | .................... | .................... | .................... |
| D | 2484 hCoV-19/Russia/Moscow_PMVL-4/2020\|EPI_ISL_470898\|2 | .................... | .................... | ..................... | .................... | .................... | .................... |
| D | 2485 hCoV-19/Russia/Moscow_PMVL-5/2020\|EPI_ISL_470899\|2 | .................... | .................... | ..................... | .................... | .................... | .................... |
| D | 2486 hCoV-19/Wales/PHWC-162663/2020\|EPI_ISL_473102\|2020 | .................... | .................... | ..................... | .................... | .................... | .................... |
| D | 2487 hCoV-19/Wales/PHWC-1626AF/2020\|EPI_ISL_473104\|2020 | .................... | .................... | ..................... | .................... | .................... | .................... |
| D | 2488 hCoV-19/Wales/PHWC-1626BE/2020\|EPI_ISL_473105\|2020 | .................... | .................... | ..................... | .................... | .................... | .................... |
| D | 2489 hCoV-19/Wales/PHWC-1626DC/2020\|EPI_ISL_473107\|2020 | .................... | .................... | ..................... | .................... | .................... | .................... |
| D | 2490 hCoV-19/Wales/PHWC-162751/2020\|EPI_ISL_473115\|2020 | .................... | .................... | ..................... | .................... | .................... | .................... |

| | | | | | | | | |
|---|---|---|---|---|---|---|---|---|
| D | 2491 | hCoV-19/Wales/PHWC-1627AC/2020\|EPI_ISL_473119\|2020 | ···················· | ···················· | ···················· | ···················· | ···················· | ···················· |
| D | 2492 | hCoV-19/Wales/PHWC-1627D9/2020\|EPI_ISL_473122\|2020 | ···················· | ···················· | ···················· | ···················· | ···················· | ···················· |
| D | 2493 | hCoV-19/Wales/PHWC-338EE/2020\|EPI_ISL_474403\|2020- | ···················· | ···················· | ···················· | ···················· | ···················· | ···················· |
| D | 2494 | hCoV-19/Wales/PHWC-34CC3/2020\|EPI_ISL_474421\|2020- | ···················· | ···················· | ···················· | ···················· | ···················· | ···················· |
| D | 2495 | hCoV-19/USA/CA-CSMC84/2020\|EPI_ISL_475700\|2020-03- | ···················· | ···················· | ···················· | ···················· | ···················· | ···················· |
| D | 2496 | hCoV-19/USA/CA-CSMC85/2020\|EPI_ISL_475701\|2020-04- | ···················· | ···················· | ···················· | ···················· | ···················· | ···················· |
| D | 2497 | hCoV-19/USA/CA-CSMC86/2020\|EPI_ISL_475702\|2020-04- | ···················· | ···················· | ···················· | ···················· | ···················· | ···················· |
| D | 2498 | hCoV-19/USA/CA-CSMC87/2020\|EPI_ISL_475703\|2020-03- | ···················· | ···················· | ···················· | ···················· | ···················· | ···················· |
| D | 2499 | hCoV-19/USA/CA-CSMC89/2020\|EPI_ISL_475705\|2020-03- | ···················· | ···················· | ···················· | ···················· | ···················· | ···················· |
| D | 2500 | hCoV-19/USA/CA-CSMC9/2020\|EPI_ISL_475706\|2020-03-2 | ···················· | ···················· | ···················· | ···················· | ···················· | ···················· |
| D | 2501 | hCoV-19/USA/CA-CSMC91/2020\|EPI_ISL_475708\|2020-03- | ···················· | ···················· | ···················· | ···················· | ···················· | ···················· |
| D | 2502 | hCoV-19/USA/CA-CSMC92/2020\|EPI_ISL_475709\|2020-04- | ···················· | ···················· | ···················· | ···················· | ···················· | ···················· |
| D | 2503 | hCoV-19/USA/CA-CSMC93/2020\|EPI_ISL_475710\|2020-04- | ···················· | ···················· | ···················· | ···················· | ···················· | ···················· |
| D | 2504 | hCoV-19/USA/CA-CSMC94/2020\|EPI_ISL_475711\|2020-04- | ···················· | ···················· | ···················· | ···················· | ···················· | ···················· |
| D | 2505 | hCoV-19/USA/CA-CSMC95/2020\|EPI_ISL_475712\|2020-04- | ···················· | ···················· | ···················· | ···················· | ···················· | ···················· |
| D | 2506 | hCoV-19/USA/CA-CSMC96/2020\|EPI_ISL_475713\|2020-04- | ···················· | ···················· | ···················· | ···················· | ···················· | ···················· |
| D | 2507 | hCoV-19/USA/CA-CSMC97/2020\|EPI_ISL_475714\|2020-04- | ···················· | ···················· | ···················· | ···················· | ···················· | ···················· |
| D | 2508 | hCoV-19/USA/CA-CSMC98/2020\|EPI_ISL_475715\|2020-04- | ···················· | ···················· | ···················· | ···················· | ···················· | ···················· |
| D | 2509 | hCoV-19/USA/CA-CSMC99/2020\|EPI_ISL_475716\|2020-03- | ···················· | ···················· | ···················· | ···················· | ···················· | ···················· |
| D | 2510 | hCoV-19/Spain/PV-HUD-70065612/2020\|EPI_ISL_475717\| | ···················· | ···················· | ···················· | ···················· | ···················· | ···················· |
| D | 2511 | hCoV-19/Spain/PV-HUD-65067673/2020\|EPI_ISL_475718\| | ···················· | ···················· | ···················· | ···················· | ···················· | ···················· |
| D | 2512 | hCoV-19/Spain/PV-HUD-70063452/2020\|EPI_ISL_475719\| | ···················· | ···················· | ···················· | ···················· | ···················· | ···················· |
| D | 2513 | hCoV-19/Spain/PV-HUD-96195073/2020\|EPI_ISL_475720\| | ···················· | ···················· | ···················· | ···················· | ···················· | ···················· |
| D | 2514 | hCoV-19/Spain/PV-HUD-98203801/2020\|EPI_ISL_475721\| | ···················· | ···················· | ···················· | ···················· | ···················· | ···················· |
| D | 2515 | hCoV-19/Egypt/CUNCI-HGC6I016/2020\|EPI_ISL_475723\|2 | ···················· | ·A·················· | ···················· | ···················· | ···················· | ··A················· |
| D | 2516 | hCoV-19/Egypt/CUNCI-HGC6I031/2020\|EPI_ISL_475724\|2 | ···················· | ·········A·········· | ···················· | ···················· | ···················· | ···················· |
| D | 2517 | hCoV-19/USA/UT-04032/2020\|EPI_ISL_475726\|2020-05-1 | ···················· | ···················· | ···················· | ···················· | ···················· | ···················· |
| D | 2518 | hCoV-19/USA/UT-04033/2020\|EPI_ISL_475727\|2020-05-1 | ···················· | ···················· | ···················· | ···················· | ···················· | ···················· |
| D | 2519 | hCoV-19/USA/UT-04034/2020\|EPI_ISL_475728\|2020-05-1 | ···················· | ···················· | ···················· | ···················· | ···················· | ···················· |
| D | 2520 | hCoV-19/USA/UT-03874/2020\|EPI_ISL_475730\|2020-05-1 | ···················· | ···················· | ···················· | ···················· | ···················· | ···················· |
| D | 2522 | hCoV-19/Egypt/MASRI-11/2020\|EPI_ISL_475753\|2020-05 | ···················· | ·A·················· | ···················· | ···················· | ···················· | ··A················· |
| D | 2523 | hCoV-19/Bangladesh/BCSIR-NILMRC-025/2020\|EPI_ISL_4 | ···················· | ···················· | ···················· | ···················· | ···················· | ···················· |
| D | 2524 | hCoV-19/Bangladesh/BCSIR-NILMRC-113/2020\|EPI_ISL_4 | ···················· | ···················· | ···················· | ···················· | ···················· | ···················· |
| D | 2525 | hCoV-19/Bangladesh/BCSIR-NILMRC-108/2020\|EPI_ISL_4 | ···················· | ···················· | ···················· | ···················· | ···················· | ···················· |
| D | 2526 | hCoV-19/Bangladesh/BCSIR-NILMRC-105/2020\|EPI_ISL_4 | ···················· | ···················· | ···················· | ···················· | ···················· | ···················· |
| D | 2527 | hCoV-19/Bangladesh/BCSIR-NILMRC-115/2020\|EPI_ISL_4 | ···················· | ···················· | ···················· | ···················· | ···················· | ···················· |
| D | 2528 | hCoV-19/Bangladesh/BCSIR-NILMRC-116/2020\|EPI_ISL_4 | ···················· | ···················· | ···················· | ···················· | ···················· | ···················· |
| D | 2529 | hCoV-19/Bangladesh/BCSIR-NILMRC-021/2020\|EPI_ISL_4 | ···················· | ···················· | ···················· | ···················· | ···················· | ···················· |
| D | 2530 | hCoV-19/Bangladesh/BCSIR-NILMRC-112/2020\|EPI_ISL_4 | ···················· | ···················· | ···················· | ···················· | ···················· | ···················· |
| D | 2531 | hCoV-19/USA/OK_08/2020\|EPI_ISL_475762\|2020 | ···················· | ···················· | ···················· | ···················· | ···················· | ···················· |
| D | 2532 | hCoV-19/Austria/CeMM0059/2020\|EPI_ISL_475766\|2020- | ···················· | ···················· | ···················· | ···················· | ···················· | ···················· |
| D | 2534 | hCoV-19/Austria/CeMM0187/2020\|EPI_ISL_475770\|2020- | ···················· | ···················· | ···················· | ···················· | ··············N····· | ···················· |
| D | 2536 | hCoV-19/Austria/CeMM0236/2020\|EPI_ISL_475772\|2020- | ···················· | ···················· | ···················· | ···················· | ···················· | ···················· |
| D | 2537 | hCoV-19/Austria/CeMM0268/2020\|EPI_ISL_475773\|2020- | ···················· | ···················· | ···················· | ···················· | ···················· | ···················· |
| D | 2538 | hCoV-19/Austria/CeMM0298/2020\|EPI_ISL_475774\|2020- | ···················· | ···················· | ···················· | ···················· | ···················· | ···················· |
| D | 2539 | hCoV-19/Austria/CeMM0299/2020\|EPI_ISL_475775\|2020- | ···················· | ···················· | ···················· | ···················· | ···················· | ···················· |
| D | 2540 | hCoV-19/Austria/CeMM0300/2020\|EPI_ISL_475776\|2020- | ···················· | ···················· | ···················· | ···················· | ···················· | ···················· |
| D | 2541 | hCoV-19/Austria/CeMM0304/2020\|EPI_ISL_475779\|2020- | ···················· | ···················· | ···················· | ···················· | ···················· | ···················· |
| D | 2542 | hCoV-19/Austria/CeMM0305/2020\|EPI_ISL_475780\|2020- | ···················· | ···················· | ···················· | ···················· | ···················· | ···················· |
| D | 2543 | hCoV-19/Austria/CeMM0307/2020\|EPI_ISL_475781\|2020- | ···················· | ···················· | ···················· | ···················· | ···················· | ···················· |
| D | 2544 | hCoV-19/Austria/CeMM0308/2020\|EPI_ISL_475782\|2020- | ···················· | ···················· | ···················· | ···················· | ···················· | ···················· |
| D | 2545 | hCoV-19/Austria/CeMM0311/2020\|EPI_ISL_475783\|2020- | ···················· | ···················· | ···················· | ···················· | ···················· | ···················· |
| D | 2546 | hCoV-19/Austria/CeMM0312/2020\|EPI_ISL_475784\|2020- | ···················· | ···················· | ···················· | ···················· | ···················· | ···················· |
| D | 2547 | hCoV-19/England/SHEF-C9926/2020\|EPI_ISL_479236\|202 | ···················· | ···················· | ···················· | ···················· | ···················· | ···················· |
| D | 2548 | hCoV-19/England/SHEF-CA71C/2020\|EPI_ISL_479242\|202 | ···················· | ···················· | ···················· | ···················· | ···················· | ···················· |
| D | 2549 | hCoV-19/England/SHEF-CB64B/2020\|EPI_ISL_479248\|202 | ···················· | ···················· | ···················· | ···················· | ···················· | ···················· |
| D | 2550 | hCoV-19/England/SHEF-CD649/2020\|EPI_ISL_479258\|202 | ···················· | ···················· | ···················· | ···················· | ···················· | ···················· |
| D | 2551 | hCoV-19/England/SHEF-CFD24/2020\|EPI_ISL_479266\|202 | ···················· | ···················· | ···················· | ···················· | ···················· | ···················· |
| D | 2552 | hCoV-19/England/SHEF-D00CD/2020\|EPI_ISL_479267\|202 | ···················· | ···················· | ···················· | ···················· | ···················· | ···················· |
| D | 2553 | hCoV-19/England/SHEF-D03B5/2020\|EPI_ISL_479270\|202 | ···················· | ···················· | ···················· | ···················· | ···················· | ···················· |
| D | 2554 | hCoV-19/England/SHEF-D14EE/2020\|EPI_ISL_479275\|202 | ···················· | ···················· | ···················· | ···················· | ···················· | ···················· |
| D | 2555 | hCoV-19/England/SHEF-D1509/2020\|EPI_ISL_479276\|202 | ···················· | ···················· | ···················· | ···················· | ···················· | ···················· |
| D | 2556 | hCoV-19/England/SHEF-D283C/2020\|EPI_ISL_479279\|202 | ···················· | ···················· | ···················· | ···················· | ···················· | ···················· |
| D | 2557 | hCoV-19/England/SHEF-D2A09/2020\|EPI_ISL_479280\|202 | ···················· | ···················· | ···················· | ···················· | ···················· | ···················· |
| D | 2558 | hCoV-19/England/SHEF-D321F/2020\|EPI_ISL_479283\|202 | ···················· | ···················· | ···················· | ···················· | ···················· | ···················· |
| D | 2559 | hCoV-19/Wales/PHWC-1605A4/2020\|EPI_ISL_479294\|2020 | ···················· | ···················· | ···················· | ···················· | ···················· | ···················· |
| D | 2560 | hCoV-19/Wales/PHWC-1607CC/2020\|EPI_ISL_479297\|2020 | ···················· | ···················· | ···················· | ···················· | ···················· | ···················· |
| D | 2561 | hCoV-19/England/20169018304/2020\|EPI_ISL_465202\|20 | ···················· | ···················· | ···················· | ···················· | ···················· | ···················· |
| D | 2562 | hCoV-19/England/20169019704/2020\|EPI_ISL_465206\|20 | ···················· | ···················· | ···················· | ···················· | ···················· | ···················· |
| D | 2563 | hCoV-19/England/20169020604/2020\|EPI_ISL_465208\|20 | ···················· | ···················· | ···················· | ···················· | ·········N·········· | ···················· |
| D | 2565 | hCoV-19/England/20169021104/2020\|EPI_ISL_465211\|20 | ···················· | ···················· | ···················· | ···················· | ···················· | ···················· |
| D | 2566 | hCoV-19/England/20169022304/2020\|EPI_ISL_465213\|20 | ···················· | ···················· | ···················· | ···················· | ···················· | ···················· |
| D | 2567 | hCoV-19/England/20169023204/2020\|EPI_ISL_465218\|20 | ···················· | ···················· | ···················· | ···················· | ···················· | ···················· |
| D | 2568 | hCoV-19/England/20169029504/2020\|EPI_ISL_465222\|20 | ···················· | ···················· | ···················· | ···················· | ···················· | ···················· |
| D | 2569 | hCoV-19/England/20169030004/2020\|EPI_ISL_465224\|20 | ···················· | ·R·················· | ···················· | ···················· | ···················· | ···················· |
| D | 2570 | hCoV-19/England/20169030304/2020\|EPI_ISL_465225\|20 | ···················· | ···················· | ···················· | ···················· | ···················· | ···················· |
| D | 2571 | hCoV-19/England/20169032504/2020\|EPI_ISL_465227\|20 | ···················· | ···················· | ···················· | ···················· | ···················· | ···················· |
| D | 2572 | hCoV-19/England/20169036204/2020\|EPI_ISL_465241\|20 | ···················· | ···················· | ···················· | ···················· | ···················· | ···················· |

| | | | | | | | | | |
|---|---|---|---|---|---|---|---|---|---|
| D | 2573 | hCoV-19/England/20169036604/2020\|EPI_ISL_465243\|20 | ···················· | ···················· | ···················· | ···················· | ···················· | ···················· | |
| D | 2574 | hCoV-19/England/20169037404/2020\|EPI_ISL_465244\|20 | ···················· | ···················· | ···················· | ···················· | ···················· | ···················· | |
| D | 2576 | hCoV-19/England/20169040504/2020\|EPI_ISL_465248\|20 | ···················· | ···················· | ···················· | ···················· | ···················· | ····N····N·········· | |
| D | 2577 | hCoV-19/England/20169042204/2020\|EPI_ISL_465253\|20 | ···················· | ···················· | ···················· | ···················· | ···················· | ···················· | |
| D | 2579 | hCoV-19/England/20169043104/2020\|EPI_ISL_465259\|20 | ···················· | ···················· | ···················· | ···················· | ···················· | ···················· | |
| D | 2580 | hCoV-19/England/20170000304/2020\|EPI_ISL_465260\|20 | ···················· | ···················· | ···················· | ···················· | ···················· | ···················· | |
| D | 2582 | hCoV-19/England/20170000704/2020\|EPI_ISL_465262\|20 | ···················· | ···················· | ···················· | ···················· | ···················· | ···················· | |
| D | 2584 | hCoV-19/England/20170002404/2020\|EPI_ISL_465266\|20 | ···················· | ···················· | ···················· | ···················· | ···················· | ···················· | |
| D | 2585 | hCoV-19/England/20166006604/2020\|EPI_ISL_466514\|20 | ···················· | ···················· | ···················· | ···················· | ···················· | ···················· | |
| D | 2586 | hCoV-19/England/20166006904/2020\|EPI_ISL_466516\|20 | ···················· | ···················· | ···················· | ···················· | ···················· | ···················· | |
| D | 2587 | hCoV-19/England/20166008604/2020\|EPI_ISL_466520\|20 | ···················· | ···················· | ···················· | ···················· | ···················· | ···················· | |
| D | 2588 | hCoV-19/England/20166010604/2020\|EPI_ISL_466525\|20 | ···················· | ···················· | ···················· | ·········K·········· | ···················· | ···················· | |
| D | 2589 | hCoV-19/England/20166012304/2020\|EPI_ISL_466530\|20 | ···················· | ···················· | ···················· | ···················· | ···················· | ···················· | |
| D | 2590 | hCoV-19/England/20166012904/2020\|EPI_ISL_466532\|20 | ···················· | ···················· | ···················· | ···················· | ···················· | ···················· | |
| D | 2591 | hCoV-19/England/20166013104/2020\|EPI_ISL_466533\|20 | ···················· | ···················· | ···················· | ···················· | ···················· | ···················· | |
| D | 2593 | hCoV-19/England/20166015304/2020\|EPI_ISL_466537\|20 | ···················· | ···················· | ···················· | ···················· | ···················· | ···················· | |
| D | 2594 | hCoV-19/England/20166015404/2020\|EPI_ISL_466538\|20 | ···················· | ···················· | ···················· | ···················· | ···················· | ···················· | |
| D | 2595 | hCoV-19/England/20166016004/2020\|EPI_ISL_466539\|20 | ···················· | ···················· | ···················· | ···················· | ···················· | ···················· | |
| D | 2596 | hCoV-19/England/20166017804/2020\|EPI_ISL_466543\|20 | ···················· | ···················· | ···················· | ···················· | ···················· | ···················· | |
| D | 2597 | hCoV-19/England/20166054804/2020\|EPI_ISL_466548\|20 | ···················· | ···················· | ···················· | ···················· | ···················· | ···················· | |
| D | 2598 | hCoV-19/England/20166055904/2020\|EPI_ISL_466550\|20 | ···················· | ···················· | ···················· | ···················· | ···················· | ···················· | |
| D | 2599 | hCoV-19/England/20166056304/2020\|EPI_ISL_466552\|20 | ···················· | ···················· | ···················· | ···················· | ···················· | ···················· | |
| D | 2601 | hCoV-19/England/20166058904/2020\|EPI_ISL_466556\|20 | ···················· | ···················· | ···················· | ···················· | ···················· | ···················· | |
| D | 2602 | hCoV-19/England/20166059104/2020\|EPI_ISL_466557\|20 | ···················· | ···················· | ···················· | ···················· | ···················· | ···················· | |
| D | 2604 | hCoV-19/England/20166060504/2020\|EPI_ISL_466559\|20 | ···················· | ···················· | ···················· | ···················· | ···················· | ···················· | |
| D | 2605 | hCoV-19/England/20166063404/2020\|EPI_ISL_466561\|20 | ···················· | ···················· | ···················· | ···················· | ···················· | ···················· | |
| D | 2606 | hCoV-19/England/20166063504/2020\|EPI_ISL_466562\|20 | ···················· | ···················· | ···················· | ···················· | ···················· | ···················· | |
| D | 2607 | hCoV-19/England/20166064004/2020\|EPI_ISL_466563\|20 | ···················· | ···················· | ···················· | ···················· | ···················· | ···················· | |
| D | 2609 | hCoV-19/England/20166067804/2020\|EPI_ISL_466569\|20 | ···················· | ···················· | ···················· | ···················· | ···················· | ···················· | |
| D | 2610 | hCoV-19/England/20166068204/2020\|EPI_ISL_466570\|20 | ···················· | ···················· | ···················· | ···················· | ···················· | ···················· | |
| D | 2611 | hCoV-19/England/20166068404/2020\|EPI_ISL_466572\|20 | ···················· | ···················· | ···················· | ···················· | ···················· | ···················· | |
| D | 2612 | hCoV-19/England/20166069104/2020\|EPI_ISL_466573\|20 | ···················· | ···················· | ···················· | ···················· | ···················· | ···················· | |
| D | 2613 | hCoV-19/England/20166069304/2020\|EPI_ISL_466574\|20 | ···················· | ···················· | ···················· | ···················· | ···················· | ···················· | |
| D | 2615 | hCoV-19/England/20166069904/2020\|EPI_ISL_466577\|20 | ···················· | ···················· | ···················· | ···················· | ···················· | ···················· | |
| D | 2616 | hCoV-19/England/20166070904/2020\|EPI_ISL_466580\|20 | ···················· | ···················· | ···················· | ···················· | ···················· | N··NN··············· | 1 |
| D | 2621 | hCoV-19/England/20168000604/2020\|EPI_ISL_466588\|20 | ···················· | ···················· | ···················· | ···················· | ···················· | ···················· | |
| D | 2624 | hCoV-19/England/20168001304/2020\|EPI_ISL_466591\|20 | ···················· | ···················· | ···················· | ···················· | ···················· | ···················· | |
| D | 2625 | hCoV-19/England/20168001704/2020\|EPI_ISL_466592\|20 | ···················· | ···················· | ···················· | ···················· | ···················· | ···················· | |
| D | 2626 | hCoV-19/England/20168002704/2020\|EPI_ISL_466593\|20 | ···················· | ···················· | ···················· | ···················· | ···················· | ···················· | |
| D | 2627 | hCoV-19/England/20168002804/2020\|EPI_ISL_466594\|20 | ···················· | ···················· | ···················· | ···················· | ···················· | ···················· | |
| D | 2628 | hCoV-19/England/20168003204/2020\|EPI_ISL_466596\|20 | ···················· | ···················· | ···················· | ···················· | ···················· | ···················· | |
| D | 2629 | hCoV-19/England/20168004104/2020\|EPI_ISL_466597\|20 | ···················· | ···················· | ···················· | ···················· | ···················· | ···················· | |
| D | 2630 | hCoV-19/England/20168005004/2020\|EPI_ISL_466598\|20 | ···················· | ···················· | ···················· | ···················· | ···················· | ···················· | |
| D | 2631 | hCoV-19/England/20168005504/2020\|EPI_ISL_466599\|20 | ···················· | ···················· | ···················· | ···················· | ···················· | ···················· | |
| D | 2632 | hCoV-19/Spain/MD-IBV-003691/2020\|EPI_ISL_467249\|20 | ···················· | ···················· | ···················· | ···················· | ···················· | ···················· | |
| D | 2637 | hCoV-19/Spain/MD-IBV-003664/2020\|EPI_ISL_467254\|20 | ···················· | ···················· | ···················· | ···················· | ···················· | ···················· | |
| D | 2638 | hCoV-19/Spain/MD-IBV-003646/2020\|EPI_ISL_467255\|20 | ···················· | ···················· | ···················· | ···················· | ···················· | ···················· | |
| D | 2639 | hCoV-19/Spain/MD-IBV-003698/2020\|EPI_ISL_467256\|20 | ···················· | ···················· | ···················· | ···················· | ···················· | ···················· | |
| D | 2640 | hCoV-19/Spain/MD-IBV-003696/2020\|EPI_ISL_467257\|20 | ···················· | ···················· | ···················· | ···················· | ···················· | ···················· | |
| D | 2641 | hCoV-19/Spain/MD-IBV-003700/2020\|EPI_ISL_467259\|20 | ···················· | ···················· | ···················· | ···················· | ···················· | ···················· | |
| D | 2642 | hCoV-19/Spain/MD-IBV-003676/2020\|EPI_ISL_467260\|20 | ···················· | ···················· | ···················· | ···················· | ···················· | ···················· | |
| D | 2643 | hCoV-19/Spain/MD-IBV-003693/2020\|EPI_ISL_467261\|20 | ···················· | ···················· | ···················· | ···················· | ···················· | ···················· | |
| D | 2645 | hCoV-19/Spain/GA-IBV-002714/2020\|EPI_ISL_467263\|20 | ···················· | ···················· | ···················· | ···················· | ···················· | ···················· | |
| D | 2648 | hCoV-19/Spain/GA-IBV-002756/2020\|EPI_ISL_467267\|20 | ···················· | ···················· | ···················· | ···················· | ···················· | ···················· | |
| D | 2654 | hCoV-19/Spain/GA-IBV-002710/2020\|EPI_ISL_467278\|20 | ···················· | ···················· | ···················· | ···················· | ···················· | ···················· | |
| D | 2656 | hCoV-19/USA/CA-QDX-73/2020\|EPI_ISL_467840\|2020-03- | ···················· | ···················· | ···················· | ···················· | ···················· | ···················· | |
| D | 2657 | hCoV-19/USA/CA-QDX-76/2020\|EPI_ISL_467841\|2020-03- | ···················· | ···················· | ···················· | ···················· | ···················· | ···················· | |
| D | 2658 | hCoV-19/USA/CA-QDX-78/2020\|EPI_ISL_467842\|2020-03- | ···················· | ···················· | ···················· | ···················· | ···················· | ···················· | |
| D | 2659 | hCoV-19/USA/CA-QDX-79/2020\|EPI_ISL_467843\|2020-03- | ···················· | ···················· | ···················· | ···················· | ···················· | ···················· | |
| D | 2660 | hCoV-19/USA/FL-QDX-121/2020\|EPI_ISL_467844\|2020-03 | ···················· | ···················· | ···················· | ···················· | ···················· | ···················· | |
| D | 2661 | hCoV-19/USA/GA-QDX-115/2020\|EPI_ISL_467845\|2020-03 | ···················· | ···················· | ···················· | ···················· | ···················· | ···················· | |
| D | 2662 | hCoV-19/USA/GA-QDX-116/2020\|EPI_ISL_467846\|2020-03 | ···················· | ···················· | ···················· | ···················· | ···················· | ···················· | |
| D | 2663 | hCoV-19/USA/GA-QDX-122/2020\|EPI_ISL_467847\|2020-03 | ···················· | ···················· | ···················· | ···················· | ···················· | ···················· | |
| D | 2664 | hCoV-19/USA/GA-QDX-152/2020\|EPI_ISL_467848\|2020-03 | ···················· | ···················· | ···················· | ···················· | ···················· | ···················· | |
| D | 2665 | hCoV-19/USA/GA-QDX-53/2020\|EPI_ISL_467849\|2020-03- | ···················· | ······A············· | ···················· | ···················· | ···················· | ···················· | |
| D | 2666 | hCoV-19/USA/MA-QDX-114/2020\|EPI_ISL_467850\|2020-03 | ···················· | ···················· | ···················· | ···················· | ···················· | ···················· | |
| D | 2667 | hCoV-19/USA/MA-QDX-127/2020\|EPI_ISL_467851\|2020-03 | ···················· | ···················· | ···················· | ···················· | ···················· | ···················· | |
| D | 2668 | hCoV-19/USA/MA-QDX-135/2020\|EPI_ISL_467852\|2020-03 | ···················· | ···················· | ···················· | ···················· | ···················· | ···················· | |
| D | 2669 | hCoV-19/USA/MA-QDX-145/2020\|EPI_ISL_467853\|2020-03 | ···················· | ···················· | ···················· | ···················· | ···················· | ···················· | |
| D | 2670 | hCoV-19/USA/MA-QDX-149/2020\|EPI_ISL_467854\|2020-03 | ···················· | ···················· | ···················· | ···················· | ···················· | ···················· | |
| D | 2671 | hCoV-19/USA/MA-QDX-150/2020\|EPI_ISL_467855\|2020-03 | ···················· | ···················· | ···················· | ···················· | ···················· | ···················· | |
| D | 2672 | hCoV-19/USA/MA-QDX-31/2020\|EPI_ISL_467856\|2020-03- | ···················· | ···················· | ···················· | ···················· | ···················· | ···················· | |
| D | 2673 | hCoV-19/USA/MA-QDX-32/2020\|EPI_ISL_467857\|2020-03- | ···················· | ···················· | ···················· | ···················· | ···················· | ···················· | |
| D | 2674 | hCoV-19/USA/MA-QDX-41/2020\|EPI_ISL_467858\|2020-03- | ···················· | ···················· | ···················· | ···················· | ···················· | ···················· | |
| D | 2675 | hCoV-19/USA/MA-QDX-47/2020\|EPI_ISL_467859\|2020-03- | ···················· | ···················· | ···················· | ···················· | ···················· | ···················· | |
| D | 2676 | hCoV-19/USA/MA-QDX-54/2020\|EPI_ISL_467860\|2020-03- | ···················· | ···················· | ···················· | ···················· | ···················· | ···················· | |
| D | 2677 | hCoV-19/USA/MA-QDX-59/2020\|EPI_ISL_467861\|2020-03- | ···················· | ···················· | ···················· | ···················· | ···················· | ···················· | |
| D | 2678 | hCoV-19/USA/MA-QDX-61/2020\|EPI_ISL_467862\|2020-03- | ···················· | ···················· | ···················· | ···················· | ···················· | ···················· | |

| | | | | | | | | |
|---|---|---|---|---|---|---|---|---|
| D | 2679 | hCoV-19/USA/MA-QDX-64/2020\|EPI_ISL_467863\|2020-03- | ···················· | ···················· | ···················· | ···················· | ···················· | ···················· |
| D | 2680 | hCoV-19/USA/MA-QDX-68/2020\|EPI_ISL_467864\|2020-03- | ···················· | ···················· | ···················· | ···················· | ···················· | ···················· |
| D | 2681 | hCoV-19/USA/MA-QDX-69/2020\|EPI_ISL_467865\|2020-03- | ···T················ | ···················· | ···················· | ···················· | ···················· | ···················· |
| D | 2682 | hCoV-19/USA/MA-QDX-80/2020\|EPI_ISL_467866\|2020-03- | ···················· | ···················· | ···················· | ···················· | ···················· | ···················· |
| D | 2683 | hCoV-19/USA/MD-QDX-113/2020\|EPI_ISL_467867\|2020-03 | ···················· | ···················· | ···················· | ···················· | ···················· | ···················· |
| D | 2684 | hCoV-19/USA/MI-QDX-26/2020\|EPI_ISL_467868\|2020-03- | ···················· | ···················· | ···················· | ···················· | ···················· | ···················· |
| D | 2685 | hCoV-19/USA/MI-QDX-28/2020\|EPI_ISL_467869\|2020-03- | ···················· | ···················· | ···················· | ···················· | ···················· | ···················· |
| D | 2686 | hCoV-19/USA/MI-QDX-52/2020\|EPI_ISL_467870\|2020-03- | ···················· | ···················· | ···················· | ···················· | ···················· | ···················· |
| D | 2687 | hCoV-19/USA/MI-QDX-58/2020\|EPI_ISL_467871\|2020-03- | ···················· | ···················· | ···················· | ···················· | ···················· | ···················· |
| D | 2688 | hCoV-19/USA/MI-QDX-70/2020\|EPI_ISL_467872\|2020-03- | ···················· | ···················· | ···················· | ···················· | ···················· | ···················· |
| D | 2689 | hCoV-19/USA/MI-QDX-71/2020\|EPI_ISL_467873\|2020-03- | ···················· | ···················· | ···················· | ···················· | ···················· | ···················· |
| D | 2690 | hCoV-19/USA/NJ-QDX-123/2020\|EPI_ISL_467874\|2020-03 | ···················· | ···················· | ···················· | ···················· | ···················· | ···················· |
| D | 2691 | hCoV-19/USA/NJ-QDX-124/2020\|EPI_ISL_467875\|2020-03 | ···················· | ···················· | ···················· | ···················· | ···················· | ···················· |
| D | 2692 | hCoV-19/USA/NJ-QDX-133/2020\|EPI_ISL_467876\|2020-03 | ···················· | ···················· | ···················· | ···················· | ···················· | ···················· |
| D | 2693 | hCoV-19/USA/NJ-QDX-141/2020\|EPI_ISL_467877\|2020-03 | ···················· | ···················· | ···················· | ···················· | ···················· | ···················· |
| D | 2694 | hCoV-19/USA/NJ-QDX-144/2020\|EPI_ISL_467878\|2020-03 | ···················· | ···················· | ···················· | ···················· | ···················· | ···················· |
| D | 2695 | hCoV-19/USA/NJ-QDX-23/2020\|EPI_ISL_467879\|2020-03- | ···················· | ···················· | ···················· | ···················· | ···················· | ···················· |
| D | 2696 | hCoV-19/USA/NJ-QDX-39/2020\|EPI_ISL_467880\|2020-03- | ···················· | ···················· | ···················· | ···················· | ···················· | ···················· |
| D | 2697 | hCoV-19/USA/NJ-QDX-55/2020\|EPI_ISL_467881\|2020-03- | ···················· | ···················· | ···················· | ···················· | ···················· | ···················· |
| D | 2698 | hCoV-19/USA/NJ-QDX-56/2020\|EPI_ISL_467882\|2020-03- | ···················· | ···················· | ···················· | ···················· | ···················· | ···················· |
| D | 2699 | hCoV-19/USA/NJ-QDX-74/2020\|EPI_ISL_467883\|2020-03- | ···················· | ···················· | ···················· | ···················· | ···················· | ···················· |
| D | 2700 | hCoV-19/USA/NY-QDX-112/2020\|EPI_ISL_467884\|2020-03 | ···················· | ···················· | ···················· | ···················· | ···················· | ···················· |
| D | 2701 | hCoV-19/USA/NY-QDX-125/2020\|EPI_ISL_467885\|2020-03 | ···················· | ···················· | ···················· | ···················· | ···················· | ···················· |
| D | 2702 | hCoV-19/USA/NY-QDX-128/2020\|EPI_ISL_467886\|2020-03 | ···················· | ···················· | ···················· | ···················· | ···················· | ···················· |
| D | 2703 | hCoV-19/USA/NY-QDX-130/2020\|EPI_ISL_467887\|2020-03 | ···················· | ···················· | ···················· | ···················· | ···················· | ···················· |
| D | 2704 | hCoV-19/USA/NY-QDX-134/2020\|EPI_ISL_467888\|2020-03 | ···················· | ···················· | ···················· | ···················· | ···················· | ···················· |
| D | 2705 | hCoV-19/USA/NY-QDX-136/2020\|EPI_ISL_467889\|2020-03 | ···················· | ···················· | ···················· | ···················· | ···················· | ···················· |
| D | 2706 | hCoV-19/USA/NY-QDX-137/2020\|EPI_ISL_467890\|2020-03 | ···················· | ···················· | ···················· | ···················· | ···················· | ···················· |
| D | 2707 | hCoV-19/USA/NY-QDX-139/2020\|EPI_ISL_467891\|2020-03 | ···················· | ···················· | ···················· | ···················· | ···················· | ···················· |
| D | 2708 | hCoV-19/USA/NY-QDX-140/2020\|EPI_ISL_467892\|2020-03 | ···················· | ···················· | ···················· | ···················· | ···················· | ···················· |
| D | 2709 | hCoV-19/USA/NY-QDX-142/2020\|EPI_ISL_467893\|2020-03 | ···················· | ···················· | ···················· | ···················· | ···················· | ···················· |
| D | 2710 | hCoV-19/USA/NY-QDX-143/2020\|EPI_ISL_467894\|2020-03 | ···················· | ···················· | ···················· | ···················· | ···················· | ···················· |
| D | 2711 | hCoV-19/USA/NY-QDX-146/2020\|EPI_ISL_467895\|2020-03 | ···················· | ···················· | ···················· | ···················· | ···················· | ···················· |
| D | 2712 | hCoV-19/USA/NY-QDX-147/2020\|EPI_ISL_467896\|2020-03 | ···················· | ···················· | ···················· | ···················· | ···················· | ···················· |
| D | 2713 | hCoV-19/USA/NY-QDX-19/2020\|EPI_ISL_467897\|2020-03- | ···················· | ···················· | ···················· | ···················· | ···················· | ···················· |
| D | 2714 | hCoV-19/USA/NY-QDX-24/2020\|EPI_ISL_467898\|2020-03- | ···················· | ···················· | ···················· | ···················· | ···················· | ···················· |
| D | 2715 | hCoV-19/USA/NY-QDX-35/2020\|EPI_ISL_467899\|2020-03- | ···················· | ···················· | ···················· | ···················· | ···················· | ···················· |
| D | 2716 | hCoV-19/USA/NY-QDX-108/2020\|EPI_ISL_468580\|2020-03 | ···················· | ···················· | ···················· | ···················· | ···················· | ···················· |
| D | 2717 | hCoV-19/USA/NY-QDX-109/2020\|EPI_ISL_468581\|2020-03 | ···················· | ···················· | ···················· | ···················· | ···················· | ···················· |
| D | 2718 | hCoV-19/USA/NY-QDX-87/2020\|EPI_ISL_468582\|2020-03- | ···················· | ···················· | ···················· | ···················· | ···················· | ···················· |
| D | 2719 | hCoV-19/USA/NY-QDX-88/2020\|EPI_ISL_468583\|2020-03- | ···················· | ···················· | ···················· | ···················· | ···················· | ···················· |
| D | 2720 | hCoV-19/USA/NY-QDX-90/2020\|EPI_ISL_468584\|2020-03- | ···················· | ···················· | ···················· | ···················· | ···················· | ···················· |
| D | 2721 | hCoV-19/USA/NY-QDX-91/2020\|EPI_ISL_468585\|2020-03- | ···················· | ·A·················· | ···················· | ···················· | ···················· | ···················· |
| D | 2722 | hCoV-19/USA/NY-QDX-94/2020\|EPI_ISL_468586\|2020-03- | ···················· | ···················· | ···················· | ···················· | ···················· | ···················· |
| D | 2723 | hCoV-19/USA/NY-QDX-95/2020\|EPI_ISL_468587\|2020-03- | ···················· | ···················· | ···················· | ···················· | ···················· | ···················· |
| D | 2724 | hCoV-19/USA/NY-QDX-96/2020\|EPI_ISL_468588\|2020-03- | ···················· | ···················· | ···················· | ···················· | ···················· | ···················· |
| D | 2725 | hCoV-19/USA/OK-QDX-93/2020\|EPI_ISL_468589\|2020-03- | ···················· | ···················· | ···················· | ···················· | ···················· | ···················· |
| D | 2726 | hCoV-19/USA/WA-QDX-104/2020\|EPI_ISL_468590\|2020-03 | ···················· | ···················· | ···················· | ···················· | ···················· | ···················· |
| D | 2727 | hCoV-19/Croatia/LG-S2new/2020\|EPI_ISL_468591\|2020- | ···················· | ···················· | ···················· | ···················· | ···················· | ···················· |
| D | 2728 | hCoV-19/USA/CA-CZB-1648/2020\|EPI_ISL_468592\|2020-0 | ···················· | ···················· | ···················· | ···················· | ···················· | ···················· |
| D | 2729 | hCoV-19/USA/CA-CZB-1649/2020\|EPI_ISL_468593\|2020-0 | ···················· | ···················· | ···················· | ···················· | ···················· | ···················· |
| D | 2730 | hCoV-19/USA/CA-CZB-1650/2020\|EPI_ISL_468594\|2020-0 | ···················· | ···················· | ···················· | ···················· | ···················· | ···················· |
| D | 2731 | hCoV-19/USA/CA-CZB-1651/2020\|EPI_ISL_468595\|2020-0 | ···················· | ···················· | ···················· | ···················· | ···················· | ···················· |
| D | 2732 | hCoV-19/USA/CA-CZB-1652/2020\|EPI_ISL_468596\|2020-0 | ···················· | ···················· | ···················· | ···················· | ···················· | ···················· |
| D | 2733 | hCoV-19/USA/CA-CZB-1653/2020\|EPI_ISL_468597\|2020-0 | ···················· | ···················· | ···················· | ···················· | ···················· | ···················· |
| D | 2734 | hCoV-19/USA/CA-CZB-1654/2020\|EPI_ISL_468598\|2020-0 | ···················· | ···················· | ···················· | ···················· | ···················· | ···················· |
| D | 2735 | hCoV-19/USA/CA-CZB-1655/2020\|EPI_ISL_468599\|2020-0 | ···················· | ···················· | ···················· | ···················· | ···················· | ···················· |
| D | 2736 | hCoV-19/Australia/WA48/2020\|EPI_ISL_470865\|2020-03 | ···················· | ···················· | ···················· | ···················· | ···················· | ···················· |
| D | 2737 | hCoV-19/Australia/WA50/2020\|EPI_ISL_470867\|2020-03 | ···················· | ···················· | ···················· | ···················· | ···················· | ···················· |
| D | 2738 | hCoV-19/Australia/WA51/2020\|EPI_ISL_470868\|2020-03 | ···················· | ···················· | ···················· | ···················· | ···················· | ···················· |
| D | 2739 | hCoV-19/Australia/WA52/2020\|EPI_ISL_470869\|2020-03 | ···················· | ···················· | ···················· | ···················· | ···················· | ···················· |
| D | 2740 | hCoV-19/Australia/WA55/2020\|EPI_ISL_470872\|2020-03 | ···················· | ···················· | ···················· | ···················· | ···················· | ···················· |
| D | 2741 | hCoV-19/Australia/WA56/2020\|EPI_ISL_470873\|2020-03 | ···················· | ···················· | ···················· | ···················· | ···················· | ···················· |
| D | 2742 | hCoV-19/Georgia/Tb-22208/2020\|EPI_ISL_470876\|2020- | ···················· | ···················· | ···················· | ···················· | ···················· | ···················· |
| D | 2743 | hCoV-19/Georgia/Tb-3118/2020\|EPI_ISL_470877\|2020-0 | ···················· | ···················· | ···················· | ···················· | ···················· | ···················· |
| D | 2744 | hCoV-19/Norway/1949/2020\|EPI_ISL_470882\|2020-03-09 | ···················· | ···················· | ···················· | ···················· | ···················· | ···················· |
| D | 2747 | hCoV-19/England/NOTT-111D04/2020\|EPI_ISL_472398\|20 | ···················· | ···················· | ···················· | ···················· | ···················· | ···················· |
| D | 2748 | hCoV-19/England/NOTT-111D13/2020\|EPI_ISL_472399\|20 | ···················· | ···················· | ···················· | ···················· | ···················· | ···················· |
| D | 2751 | hCoV-19/Scotland/CVR3554/2020\|EPI_ISL_473689\|2020- | ···················· | ·A·················· | ···················· | ···················· | ···················· | ···················· |
| D | 2752 | hCoV-19/Italy/BAS-UniMI-2161/2020\|EPI_ISL_477200\|2 | ···················· | ···················· | ···················· | ···················· | ···················· | ···················· |
| D | 2753 | hCoV-19/Italy/APU-UniMI-694/2020\|EPI_ISL_477203\|20 | ···················· | ···················· | ···················· | ···················· | ···················· | ···················· |
| D | 2754 | hCoV-19/India/KA-InStem-NCBS-0002/2020\|EPI_ISL_477 | ···················· | ···················· | ···················· | ···················· | ···················· | ···················· |
| D | 2755 | hCoV-19/India/KA-InStem-NCBS-0004/2020\|EPI_ISL_477 | ···················· | ···················· | ···················· | ···················· | ···················· | ···················· |
| D | 2756 | hCoV-19/India/KA-InStem-NCBS-0007/2020\|EPI_ISL_477 | ···················· | ·A·················· | ···················· | ···················· | ···················· | ···A················ |
| D | 2757 | hCoV-19/India/KA-InStem-NCBS-0008/2020\|EPI_ISL_477 | ···················· | ·A·················· | ···················· | ···················· | ···················· | ···A················ |
| D | 2758 | hCoV-19/India/KA-InStem-NCBS-0038/2020\|EPI_ISL_477 | ···················· | ·A·················· | ···················· | ···················· | ···················· | ···A················ |
| D | 2759 | hCoV-19/India/KA-InStem-NCBS-0040/2020\|EPI_ISL_477 | ···················· | ···················· | ···················· | ···················· | ···················· | ···················· |
| D | 2760 | hCoV-19/India/KA-InStem-NCBS-0041/2020\|EPI_ISL_477 | ···················· | ···················· | ···················· | ···················· | ···················· | ···················· |

| | | | | | | | | |
|---|---|---|---|---|---|---|---|---|
| D | 2761 hCoV-19/India/KA-InStem-NCBS-0046/2020\|EPI_ISL_477 | ···················· | ···················· | ···················· | ···················· | ···················· | ···················· | |
| D | 2762 hCoV-19/England/NORT-283971/2020\|EPI_ISL_478513\|20 | ···················· | ···················· | ···················· | ···················· | ···················· | ···················· | |
| D | 2763 hCoV-19/England/NORT-283A23/2020\|EPI_ISL_478515\|20 | ···················· | ···················· | ···················· | ···················· | ···················· | ···················· | |
| D | 2764 hCoV-19/England/NORT-283A8D/2020\|EPI_ISL_478516\|20 | ···················· | ···················· | ···················· | ···················· | ···················· | ···················· | |
| D | 2765 hCoV-19/England/NORT-283A9C/2020\|EPI_ISL_478517\|20 | ···················· | ···················· | ···················· | ···················· | ···················· | ···················· | |
| D | 2766 hCoV-19/England/NORT-283B11/2020\|EPI_ISL_478519\|20 | ···················· | ···················· | ···················· | ···················· | ···················· | ···················· | |
| D | 2767 hCoV-19/England/NORT-283BF3/2020\|EPI_ISL_478520\|20 | ···················· | ···················· | ···················· | ···················· | ···················· | ···················· | |
| D | 2768 hCoV-19/England/NORT-283CD2/2020\|EPI_ISL_478521\|20 | ···················· | ···················· | ···················· | ···················· | ···················· | ···················· | |
| D | 2769 hCoV-19/England/NORT-283CE1/2020\|EPI_ISL_478522\|20 | ···················· | ···················· | ···················· | ···················· | ···················· | ···················· | |
| D | 2770 hCoV-19/England/NORT-283ECD/2020\|EPI_ISL_478524\|20 | ···················· | ···················· | ···················· | ···················· | ···················· | ···················· | |
| D | 2771 hCoV-19/England/NORT-283EFA/2020\|EPI_ISL_478525\|20 | ···················· | ···················· | ···················· | ···················· | ···················· | ···················· | |
| D | 2772 hCoV-19/England/NORT-283F51/2020\|EPI_ISL_478527\|20 | ···················· | ···················· | ···················· | ···················· | ···················· | ···················· | |
| D | 2773 hCoV-19/England/NORT-283F60/2020\|EPI_ISL_478528\|20 | ··················T· | ···················· | ···················· | ···················· | ···················· | ···················· | 1 |
| D | 2774 hCoV-19/England/NORT-283F7F/2020\|EPI_ISL_478529\|20 | ···················· | ···················· | ···················· | ···················· | ···················· | ···················· | |
| D | 2775 hCoV-19/England/NORT-283F8E/2020\|EPI_ISL_478530\|20 | ···················· | ···················· | ···················· | ···················· | ···················· | ···················· | |
| D | 2776 hCoV-19/England/NORT-284020/2020\|EPI_ISL_478531\|20 | ···················· | ···················· | ···················· | ···················· | ···················· | ···················· | |
| D | 2777 hCoV-19/England/NORT-28403F/2020\|EPI_ISL_478532\|20 | ···················· | ···················· | ···················· | ···················· | ···················· | ···················· | |
| D | 2778 hCoV-19/England/NORT-28420C/2020\|EPI_ISL_478536\|20 | ···················· | ···················· | ···················· | ···················· | ···················· | ···················· | |
| D | 2779 hCoV-19/England/NORT-284248/2020\|EPI_ISL_478537\|20 | ···················· | ···················· | ···················· | ···················· | ···················· | ···················· | |
| D | 2780 hCoV-19/England/NORT-284266/2020\|EPI_ISL_478539\|20 | ···················· | ···················· | ···················· | ···················· | ···················· | ···················· | |
| D | 2781 hCoV-19/England/NORT-284327/2020\|EPI_ISL_478541\|20 | ···················· | ···················· | ···················· | ···················· | ···················· | ···················· | |
| D | 2782 hCoV-19/England/NORT-284372/2020\|EPI_ISL_478543\|20 | ···················· | ···················· | ···················· | ···················· | ···················· | ···················· | |
| D | 2783 hCoV-19/England/NORT-284381/2020\|EPI_ISL_478544\|20 | ···················· | ···················· | ···················· | ···················· | ···················· | ···················· | |
| D | 2784 hCoV-19/England/NORT-284406/2020\|EPI_ISL_478546\|20 | ···················· | ···················· | ···················· | ···················· | ···················· | ···················· | |
| D | 2785 hCoV-19/England/NORT-284415/2020\|EPI_ISL_478547\|20 | ···················· | ···················· | ···················· | ···················· | ···················· | ···················· | |
| D | 2786 hCoV-19/England/NORT-284424/2020\|EPI_ISL_478548\|20 | ···················· | ···················· | ···················· | ···················· | ···················· | ···················· | |
| D | 2787 hCoV-19/England/NORT-284433/2020\|EPI_ISL_478549\|20 | ···················· | ···················· | ···················· | ···················· | ···················· | ···················· | |
| D | 2788 hCoV-19/England/NORT-284442/2020\|EPI_ISL_478550\|20 | ···················· | ···················A | ···················· | ···················· | ···················· | ···················· | 1 |
| D | 2789 hCoV-19/England/NORT-28449D/2020\|EPI_ISL_478551\|20 | ···················· | ···················· | ···················· | ···················· | ···················· | ···················· | |
| D | 2790 hCoV-19/England/NORT-284503/2020\|EPI_ISL_478553\|20 | ···················· | ···················· | ···················· | ···················· | ···················· | ···················· | |
| D | 2791 hCoV-19/England/NORT-284521/2020\|EPI_ISL_478555\|20 | ···················· | ···················· | ···················· | ···················· | ···················· | ···················· | |
| D | 2792 hCoV-19/England/NORT-28461F/2020\|EPI_ISL_478556\|20 | ···················· | ···················· | ···················· | ···················· | ···················· | ···················· | |
| D | 2793 hCoV-19/England/NORT-28463D/2020\|EPI_ISL_478558\|20 | ···················· | ···················· | ···················· | ···················· | ···················· | ···················· | |
| D | 2794 hCoV-19/England/NORT-28464C/2020\|EPI_ISL_478559\|20 | ···················· | ···················· | ···················· | ···················· | ···················· | ···················· | |
| D | 2795 hCoV-19/England/NORT-284697/2020\|EPI_ISL_478561\|20 | ···················· | ···················· | ···················· | ···················· | ···················· | ···················· | |
| D | 2796 hCoV-19/England/NORT-2846E2/2020\|EPI_ISL_478564\|20 | ···················· | ···················· | ···················· | ···················· | ···················· | ···················· | |
| D | 2797 hCoV-19/England/NORT-284767/2020\|EPI_ISL_478565\|20 | ···················· | ···················· | ···················· | ···················· | ···················· | ···················· | |
| D | 2798 hCoV-19/England/NORT-284785/2020\|EPI_ISL_478566\|20 | ···················· | ···················· | ···················· | ···················· | ···················· | ···················· | |
| D | 2799 hCoV-19/England/NORT-2847B2/2020\|EPI_ISL_478567\|20 | ···················· | ···················· | ···················· | ···················· | ···················· | ···················· | |
| D | 2800 hCoV-19/England/NORT-2847C1/2020\|EPI_ISL_478568\|20 | ···················· | ···················· | ···················· | ···················· | ···················· | ···················· | |
| D | 2801 hCoV-19/England/NORT-2847D0/2020\|EPI_ISL_478569\|20 | ···················· | ···················· | ···················· | ···················· | ···················· | ···················· | |
| D | 2802 hCoV-19/England/NORT-2848BF/2020\|EPI_ISL_478571\|20 | ···················· | ···················· | ···················· | ···················· | ···················· | ···················· | |
| D | 2803 hCoV-19/England/NORT-2848EC/2020\|EPI_ISL_478573\|20 | ···················· | ···················· | ···················· | ···················· | ···················· | ···················· | |
| D | 2804 hCoV-19/England/NORT-284A31/2020\|EPI_ISL_478575\|20 | ···················· | ···················· | ···················· | ···················· | ···················· | ···················· | |
| D | 2805 hCoV-19/England/NORT-284A5F/2020\|EPI_ISL_478576\|20 | ···················· | ···················· | ···················· | ···················· | ···················· | ···················· | |
| D | 2806 hCoV-19/England/NORT-284A9B/2020\|EPI_ISL_478577\|20 | ···················· | ···················· | ···················· | ···················· | ···················· | ···················· | |
| D | 2807 hCoV-19/England/NORT-284B4D/2020\|EPI_ISL_478578\|20 | ···················· | ···················· | ···················· | ···················· | ···················· | ···················· | |
| D | 2808 hCoV-19/England/SHEF-C2116/2020\|EPI_ISL_479201\|202 | ···················· | ···················· | ···················· | ···················· | ···················· | ···················· | |
| D | 2809 hCoV-19/England/SHEF-C5702/2020\|EPI_ISL_479214\|202 | ···················· | ···················· | ···················· | ···················· | ···················· | ···················· | |
| D | 2810 hCoV-19/England/SHEF-C580F/2020\|EPI_ISL_479216\|202 | ···················· | ···················· | ···················· | ···················· | ···················· | ···················· | |
| D | 2811 hCoV-19/England/SHEF-C6446/2020\|EPI_ISL_479218\|202 | ···················· | ···················· | ···················· | ···················· | ···················· | ···················· | |
| D | 2812 hCoV-19/England/SHEF-C674D/2020\|EPI_ISL_479221\|202 | ···················· | ···················· | ···················· | ···················· | ···················· | ···················· | |
| D | 2813 hCoV-19/England/SHEF-C6CB7/2020\|EPI_ISL_479223\|202 | ···················· | ···················· | ···················· | ···················· | ···················· | ···················· | |
| D | 2814 hCoV-19/England/SHEF-C7032/2020\|EPI_ISL_479224\|202 | ···················· | ···················· | ···················· | ···················· | ···················· | ···················· | |
| D | 2815 hCoV-19/England/SHEF-C72D2/2020\|EPI_ISL_479225\|202 | ···················· | ···················· | ···················· | ···················· | ···················· | ···················· | |
| D | 2816 hCoV-19/England/SHEF-C730C/2020\|EPI_ISL_479226\|202 | ···················· | ···················· | ···················· | ···················· | ···················· | ···················· | |
| D | 2817 hCoV-19/USA/MN-MDH-1055/2020\|EPI_ISL_462900\|2020-0 | ···················· | ·A·················· | ···················· | ···················· | ···················· | ···················· | |
| D | 2818 hCoV-19/USA/MN-MDH-1056/2020\|EPI_ISL_462901\|2020-0 | ···················· | ·A·················· | ···················· | ···················· | ···················· | ···················· | |
| D | 2819 hCoV-19/USA/MN-MDH-1057/2020\|EPI_ISL_462902\|2020-0 | ···················· | ·A·················· | ···················· | ···················· | ···················· | ···················· | |
| D | 2820 hCoV-19/USA/MN-MDH-1058/2020\|EPI_ISL_462903\|2020-0 | ···················· | ·A·················· | ···················· | ···················· | ···················· | ···················· | |
| D | 2821 hCoV-19/USA/MN-MDH-1059/2020\|EPI_ISL_462904\|2020-0 | ···················· | ·A·················· | ···················· | ···················· | ···················· | ···················· | |
| D | 2822 hCoV-19/USA/MN-MDH-1060/2020\|EPI_ISL_462905\|2020-0 | ···················· | ·A·················· | ···················· | ···················· | ···················· | ···················· | |
| D | 2823 hCoV-19/USA/MN-MDH-1061/2020\|EPI_ISL_462906\|2020-0 | ···················· | ·A·················· | ···················· | ···················· | ···················· | ···················· | |
| D | 2824 hCoV-19/USA/MN-MDH-1062/2020\|EPI_ISL_462907\|2020-0 | ···················· | ·A·················· | ···················· | ···················· | ···················· | ···················· | |
| D | 2825 hCoV-19/USA/MN-MDH-1063/2020\|EPI_ISL_462908\|2020-0 | ···················· | ·A·················· | ···················· | ···················· | ···················· | ···················· | |
| D | 2826 hCoV-19/USA/MN-MDH-1064/2020\|EPI_ISL_462909\|2020-0 | ···················· | ·A·················· | ···················· | ···················· | ···················· | ···················· | |
| D | 2827 hCoV-19/USA/WY-WYPHL-00060/2020\|EPI_ISL_462952\|202 | ···················· | ···················· | ···················· | ···················· | ···················· | ···················· | |
| D | 2828 hCoV-19/USA/WY-WYPHL-00064/2020\|EPI_ISL_462956\|202 | ···················· | ···················· | ···················· | ···················· | ···················· | ···················· | |
| D | 2829 hCoV-19/USA/WY-WYPHL-00065/2020\|EPI_ISL_462957\|202 | ···················· | ···················· | ···················· | ···················· | ···················· | ···················· | |
| D | 2830 hCoV-19/USA/WY-WYPHL-00066/2020\|EPI_ISL_462958\|202 | ···················· | ···················· | ···················· | ···················· | ···················· | ···················· | |
| D | 2831 hCoV-19/USA/WY-WYPHL-00073/2020\|EPI_ISL_462965\|202 | ···················· | ···················· | ···················· | ···················· | ···················· | ···················· | |
| D | 2832 hCoV-19/USA/WY-WYPHL-00074/2020\|EPI_ISL_462966\|202 | ···················· | ···················· | ···················· | ···················· | ···················· | ···················· | |
| D | 2833 hCoV-19/USA/WY-WYPHL-00077/2020\|EPI_ISL_462969\|202 | ···················· | ···················· | ···················· | ···················· | ···················· | ···················· | |
| D | 2834 hCoV-19/USA/WY-WYPHL-00084/2020\|EPI_ISL_462975\|202 | ···················· | ···················· | ···················· | ···················· | ···················· | ···················· | |
| D | 2835 hCoV-19/USA/WY-WYPHL-00085/2020\|EPI_ISL_462976\|202 | ···················· | ···················· | ···················· | ···················· | ···················· | ···················· | |
| D | 2836 hCoV-19/USA/WY-WYPHL-00086/2020\|EPI_ISL_462977\|202 | ···················· | ···················· | ···················· | ···················· | ···················· | ···················· | |
| D | 2837 hCoV-19/USA/WY-WYPHL-00088/2020\|EPI_ISL_462979\|202 | ···················· | ···················· | ···················· | ···················· | ···················· | ···················· | |
| D | 2838 hCoV-19/USA/WY-WYPHL-00091/2020\|EPI_ISL_462982\|202 | ···················· | ···················· | ···················· | ···················· | ···················· | ···················· | |

| | | | | | | | | | | |
|---|---|---|---|---|---|---|---|---|---|---|
| D | 2839 hCoV-19/USA/SC-DHEC-0002/2020\|EPI_ISL_462991\|2020- | ···················· | ···················· | ···················· | ···················· | ···················· | ···················· | | | |
| D | 2840 hCoV-19/Nigeria/NG57752/2020\|EPI_ISL_462992\|2020-0 | ···················· | ···················· | ···················· | ···················· | ···················· | ···················· | | | |
| D | 2841 hCoV-19/England/20169002204/2020\|EPI_ISL_465170\|20 | ···················· | ···················· | ···················· | ···················· | ···················· | ···················· | | | |
| D | 2842 hCoV-19/England/20169003304/2020\|EPI_ISL_465173\|20 | ···················· | ···················· | ···················· | ···················· | ···················· | ···················· | | | |
| D | 2843 hCoV-19/England/20169003404/2020\|EPI_ISL_465174\|20 | ···················· | ···················· | ···················· | ···················· | ···················· | ···················· | | | |
| D | 2844 hCoV-19/England/20169004504/2020\|EPI_ISL_465178\|20 | ···················· | ···················· | ···················· | ···················· | ···················· | ···················· | | | |
| D | 2845 hCoV-19/England/20169005204/2020\|EPI_ISL_465180\|20 | ···················· | ···················· | ···················· | ···················· | ···················· | ···················· | | | |
| D | 2849 hCoV-19/England/20169005804/2020\|EPI_ISL_465184\|20 | ···················· | ···················· | ···················· | ···················· | ···················· | ···················· | | | |
| D | 2850 hCoV-19/England/20169006104/2020\|EPI_ISL_465186\|20 | ···················· | ···················· | ···················· | ···················· | ···················· | ···················· | | | |
| D | 2851 hCoV-19/England/20166004204/2020\|EPI_ISL_466506\|20 | ···················· | ···················· | ···················· | ···················· | ···················· | ···················· | | | |
| D | 2852 hCoV-19/England/20166004304/2020\|EPI_ISL_466507\|20 | ···················· | ···················· | ···················· | ···················· | ···················· | ···················· | | | |
| D | 2853 hCoV-19/England/20166004704/2020\|EPI_ISL_466508\|20 | ···················· | ···················· | ···················· | ···················· | ···················· | ···················· | | | |
| D | 2854 hCoV-19/England/20166006104/2020\|EPI_ISL_466511\|20 | ···················· | ···················· | ···················· | ···················· | ···················· | ···················· | | | |
| D | 2855 hCoV-19/USA/VA-DCLS-0406/2020\|EPI_ISL_467800\|2020- | ···················· | ···················· | ···················· | ···················· | ···················· | ···················· | | | |
| D | 2856 hCoV-19/USA/VA-DCLS-0407/2020\|EPI_ISL_467801\|2020- | ···················· | ···················· | ···················· | ···················· | ···················· | ···················· | | | |
| D | 2857 hCoV-19/USA/VA-DCLS-0408/2020\|EPI_ISL_467802\|2020- | ···················· | ···················· | ···················· | ···················· | ···················· | ···················· | | | |
| D | 2858 hCoV-19/USA/VA-DCLS-0409/2020\|EPI_ISL_467803\|2020- | ···················· | ···················· | ···················· | ···················· | ···················· | ···················· | | | |
| D | 2859 hCoV-19/USA/VA-DCLS-0410/2020\|EPI_ISL_467804\|2020- | ···················· | ···················· | ···················· | ···················· | ···················· | ···················· | | | |
| D | 2860 hCoV-19/USA/VA-DCLS-0411/2020\|EPI_ISL_467805\|2020- | ···················· | ···················· | ···················· | ···················· | ···················· | ···················· | | | |
| D | 2861 hCoV-19/USA/VA-DCLS-0412/2020\|EPI_ISL_467806\|2020- | ···················· | ···················· | ···················· | ···················· | ···················· | ···················· | | | |
| D | 2862 hCoV-19/USA/VA-DCLS-0413/2020\|EPI_ISL_467807\|2020- | ···················· | ···················· | ···················· | ···················· | ···················· | ···················· | | | |
| D | 2863 hCoV-19/USA/VA-DCLS-0414/2020\|EPI_ISL_467808\|2020- | ···················· | ···················· | ···················· | ···················· | ···················· | ···················· | | | |
| D | 2864 hCoV-19/USA/CA-CSMC1/2020\|EPI_ISL_467809\|2020-03-2 | ···················· | ···················· | ···················· | ···················· | ···················· | ···················· | | | |
| D | 2865 hCoV-19/USA/CA-QDX-117/2020\|EPI_ISL_467811\|2020-03 | ···················· | ···················· | ···················· | ···················· | ···················· | ···················· | | | |
| D | 2866 hCoV-19/USA/CA-QDX-129/2020\|EPI_ISL_467812\|2020-03 | ···················· | ···················· | ···················· | ···················· | ···················· | ···················· | | | |
| D | 2867 hCoV-19/USA/CA-QDX-138/2020\|EPI_ISL_467813\|2020-03 | ···················· | ···················· | ···················· | ···················· | ···················· | ···················· | | | |
| D | 2868 hCoV-19/USA/CA-QDX-148/2020\|EPI_ISL_467814\|2020-03 | ···················· | ···················· | ···················· | ···················· | ···················· | ···················· | | | |
| D | 2869 hCoV-19/USA/CA-QDX-153/2020\|EPI_ISL_467815\|2020-03 | ···················· | ···················· | ···················· | ···················· | ···················· | ···················· | | | |
| D | 2870 hCoV-19/USA/CA-QDX-154/2020\|EPI_ISL_467816\|2020-03 | ···················· | ···················· | ···················· | ···················· | ···················· | ···················· | | | |
| D | 2871 hCoV-19/USA/CA-QDX-155/2020\|EPI_ISL_467817\|2020-03 | ···················· | ···················· | ···················· | ···················· | ···················· | ···················· | | | |
| D | 2872 hCoV-19/USA/CA-QDX-156/2020\|EPI_ISL_467818\|2020-03 | ···················· | ···················· | ···················· | ···················· | ···················· | ···················· | | | |
| D | 2873 hCoV-19/USA/CA-QDX-157/2020\|EPI_ISL_467819\|2020-03 | ···················· | ···················· | ···················· | ···················· | ···················· | ···················· | | | |
| D | 2874 hCoV-19/USA/CA-QDX-158/2020\|EPI_ISL_467820\|2020-03 | ···················· | ···················· | ···················· | ···················· | ···················· | ···················· | | | |
| D | 2875 hCoV-19/USA/CA-QDX-159/2020\|EPI_ISL_467821\|2020-03 | ···················· | ···················· | ···················· | ···················· | ···················· | ···················· | | | |
| D | 2876 hCoV-19/USA/CA-QDX-160/2020\|EPI_ISL_467822\|2020-03 | ···················· | ···················· | ···················· | ···················· | ···················· | ···················· | | | |
| D | 2877 hCoV-19/USA/CA-QDX-161/2020\|EPI_ISL_467823\|2020-03 | ···················· | ···················· | ···················· | ···················· | ···················· | ···················· | | | |
| D | 2878 hCoV-19/USA/CA-QDX-162/2020\|EPI_ISL_467824\|2020-03 | ···················· | ···················· | ···················· | ···················· | ···················· | ···················· | | | |
| D | 2879 hCoV-19/USA/CA-QDX-21/2020\|EPI_ISL_467825\|2020-03- | ···················· | ···················· | ···················· | ···················· | ···················· | ···················· | | | |
| D | 2880 hCoV-19/USA/CA-QDX-33/2020\|EPI_ISL_467827\|2020-03- | ···················· | ···················· | ···················· | ···················· | ···················· | ···················· | | | |
| D | 2881 hCoV-19/USA/CA-QDX-34/2020\|EPI_ISL_467828\|2020-03- | ···················· | ···················· | ···················· | ···················· | ···················· | ···················· | | | |
| D | 2882 hCoV-19/USA/CA-QDX-36/2020\|EPI_ISL_467829\|2020-03- | ···················· | ···················· | ···················· | ···················· | ···················· | ···················· | | | |
| D | 2883 hCoV-19/USA/CA-QDX-38/2020\|EPI_ISL_467830\|2020-03- | ···················· | ···················· | ···················· | ···················· | ···················· | ···················· | | | |
| D | 2884 hCoV-19/USA/CA-QDX-40/2020\|EPI_ISL_467831\|2020-03- | ···················· | ···················· | ···················· | ···················· | ···················· | ···················· | | | |
| D | 2885 hCoV-19/USA/CA-QDX-43/2020\|EPI_ISL_467832\|2020-03- | ···················· | ···················· | ···················· | ···················· | ···················· | ···················· | | | |
| D | 2886 hCoV-19/USA/CA-QDX-44/2020\|EPI_ISL_467833\|2020-03- | ···················· | ···················· | ···················· | ···················· | ···················· | ···················· | | | |
| D | 2887 hCoV-19/USA/CA-QDX-45/2020\|EPI_ISL_467834\|2020-03- | ···················· | ···················· | ···················· | ···················· | ···················· | ···················· | | | |
| D | 2888 hCoV-19/USA/CA-QDX-46/2020\|EPI_ISL_467835\|2020-03- | ···················· | ···················· | ···················· | ···················· | ···················· | ···················· | | | |
| D | 2889 hCoV-19/USA/CA-QDX-48/2020\|EPI_ISL_467836\|2020-03- | ···················· | ···················· | ···················· | ···················· | ···················· | ···················· | | | |
| D | 2890 hCoV-19/USA/CA-QDX-50/2020\|EPI_ISL_467837\|2020-03- | ···················· | ···················· | ···················· | ···················· | ···················· | ···················· | | | |
| D | 2891 hCoV-19/USA/CA-QDX-51/2020\|EPI_ISL_467838\|2020-03- | ···················· | ···················· | ···················· | ···················· | ···················· | ···················· | | | |
| D | 2892 hCoV-19/USA/CA-QDX-67/2020\|EPI_ISL_467839\|2020-03- | ···················· | ···················· | ···················· | ···················· | ···················· | ···················· | | | |
| D | 2893 hCoV-19/England/NORW-EB0ED/2020\|EPI_ISL_472300\|202 | ···················· | ···················· | ···················· | ···················· | ···················· | ···················· | | | |
| D | 2897 hCoV-19/England/NORW-EB1CC/2020\|EPI_ISL_472305\|202 | ···················· | ···················· | ···················· | ···················· | ···················· | ···················· | | | |
| D | 2898 hCoV-19/England/NORW-EB1EA/2020\|EPI_ISL_472306\|202 | ···················· | ···················· | ···················· | ···················· | ···················· | ···················· | | | |
| D | 2900 hCoV-19/England/NORW-EB6CD/2020\|EPI_ISL_472310\|202 | ···················· | ···················· | ···················· | ···················· | ···················· | ···················· | | | |
| D | 2901 hCoV-19/England/NORW-EB706/2020\|EPI_ISL_472311\|202 | ···················· | ···················· | ···················· | ···················· | ···················· | ···················· | | | |
| D | 2905 hCoV-19/England/NORW-EB7BB/2020\|EPI_ISL_472317\|202 | ···················· | ···················· | ···················· | ···················· | ···················· | ···················· | | | |
| D | 2906 hCoV-19/England/NORW-EB812/2020\|EPI_ISL_472320\|202 | ···················· | ···················· | ···················· | ···················· | NNN················· | ···················· | | 1 | |
| D | 2908 hCoV-19/England/NORW-EBF86/2020\|EPI_ISL_472326\|202 | ··················T· | ···················· | ···················· | ···················· | NNN················· | ···················· | | 1 | 1 |
| D | 2912 hCoV-19/England/NORW-ECB90/2020\|EPI_ISL_472336\|202 | ···················· | ···················· | ···················· | ···················· | ···················· | ···················· | | | |
| D | 2913 hCoV-19/England/NORW-ECD12/2020\|EPI_ISL_472342\|202 | ···················· | ···················· | ···················· | ···················· | NNN················· | ···················· | | 1 | |
| D | 2927 hCoV-19/England/NOTT-111BFB/2020\|EPI_ISL_472384\|20 | ···················· | ···················· | ···················· | ···················· | ···················· | ···················· | | | |
| D | 2928 hCoV-19/England/NOTT-111C16/2020\|EPI_ISL_472385\|20 | ···················· | ···················· | ···················· | ···················· | ···················· | ···················· | | | |
| D | 2929 hCoV-19/England/NOTT-111C25/2020\|EPI_ISL_472386\|20 | ···················· | ···················· | ···················· | ···················· | ···················· | ···················· | | | |
| D | 2930 hCoV-19/England/NOTT-111C34/2020\|EPI_ISL_472387\|20 | ···················· | ···················· | ···················· | ···················· | ···················· | ···················· | | | |
| D | 2931 hCoV-19/England/NOTT-111C43/2020\|EPI_ISL_472388\|20 | ···················· | ···················· | ···················· | ···················· | ···················· | ···················· | | | |
| D | 2932 hCoV-19/England/NOTT-111C61/2020\|EPI_ISL_472390\|20 | ···················· | ···················· | ···················· | ···················· | ···················· | ···················· | | | |
| D | 2933 hCoV-19/England/NOTT-111C8F/2020\|EPI_ISL_472392\|20 | ···················· | ···················· | ···················· | ···················· | ···················· | ···················· | | | |
| D | 2934 hCoV-19/England/NOTT-111CAD/2020\|EPI_ISL_472393\|20 | ···················· | ···················· | ···················· | ···················· | ···················· | ···················· | | | |
| D | 2935 hCoV-19/England/NOTT-111CBC/2020\|EPI_ISL_472394\|20 | ···················· | ···················· | ···················· | ···················· | ···················· | ···················· | | | |
| D | 2936 hCoV-19/England/NOTT-111CDA/2020\|EPI_ISL_472395\|20 | ···················· | ···················· | ···················· | ···················· | ···················· | ···················· | | | |
| D | 2937 hCoV-19/England/NOTT-111CE9/2020\|EPI_ISL_472396\|20 | ···················· | ···················· | ···················· | ···················· | ···················· | ···················· | | | |
| D | 2938 hCoV-19/Scotland/CVR2857/2020\|EPI_ISL_473600\|2020- | ···················· | ···················A | ···················· | ···················· | ···················· | ···················· | 1 | | |
| D | 2939 hCoV-19/Scotland/CVR2858/2020\|EPI_ISL_473601\|2020- | ···················· | ···················· | ···················· | ···················· | ···················· | ···················· | | | |
| D | 2940 hCoV-19/Scotland/CVR2861/2020\|EPI_ISL_473602\|2020- | ···················· | ···················· | ···················· | ···················· | ···················· | ···················· | | | |
| D | 2941 hCoV-19/Scotland/CVR2862/2020\|EPI_ISL_473603\|2020- | ···················· | ···················· | ···················· | ···················· | ···················· | ···················· | | | |
| D | 2942 hCoV-19/Scotland/CVR2863/2020\|EPI_ISL_473604\|2020- | ···················· | ···················· | ···················· | ···················· | ···················· | ···················· | | | |
| D | 2943 hCoV-19/Scotland/CVR2864/2020\|EPI_ISL_473605\|2020- | ···················· | ··············C····· | ···················· | ···················· | ···················· | ···················· | | | |

| | | | | | | | | | |
|---|---|---|---|---|---|---|---|---|---|
| D | 2944 | hCoV-19/Scotland/CVR2865/2020\|EPI_ISL_473606\|2020- | .................... | .................... | .................... | .................... | .................... | .................... | |
| D | 2945 | hCoV-19/Scotland/CVR2867/2020\|EPI_ISL_473608\|2020- | .................... | .................... | .................... | .................... | .................... | .................... | |
| D | 2946 | hCoV-19/Scotland/CVR2868/2020\|EPI_ISL_473609\|2020- | .................... | .................... | .................... | .................... | .................... | .................... | |
| D | 2947 | hCoV-19/Scotland/CVR2872/2020\|EPI_ISL_473610\|2020- | .................... | .................... | .................... | .................... | .................... | .................... | |
| D | 2948 | hCoV-19/Scotland/CVR2875/2020\|EPI_ISL_473612\|2020- | .................... | .................... | .................... | .................... | .................... | .................... | |
| D | 2949 | hCoV-19/Scotland/CVR2876/2020\|EPI_ISL_473613\|2020- | .................... | .................... | .................... | .................... | .................... | .................... | |
| D | 2950 | hCoV-19/Scotland/CVR2878/2020\|EPI_ISL_473614\|2020- | .................... | ...................A | .................... | .................... | .................... | .................... | 1 |
| D | 2951 | hCoV-19/Scotland/CVR2879/2020\|EPI_ISL_473615\|2020- | .................... | ...................A | .................... | .................... | .................... | .................... | 1 |
| D | 2952 | hCoV-19/Scotland/CVR2884/2020\|EPI_ISL_473616\|2020- | .................... | .................... | .................... | .................... | .................... | .................... | |
| D | 2953 | hCoV-19/Scotland/CVR2885/2020\|EPI_ISL_473617\|2020- | .................... | ...................A | .................... | .................... | .................... | .................... | 1 |
| D | 2954 | hCoV-19/Scotland/CVR2886/2020\|EPI_ISL_473618\|2020- | .................... | .................... | .................... | .................... | .................... | .................... | |
| D | 2955 | hCoV-19/Scotland/CVR2896/2020\|EPI_ISL_473619\|2020- | .................... | .................... | .................... | .................... | .................... | .................... | |
| D | 2956 | hCoV-19/Scotland/CVR2899/2020\|EPI_ISL_473621\|2020- | .................... | .................... | .................... | .................... | .................... | .................... | |
| D | 2957 | hCoV-19/Scotland/CVR2901/2020\|EPI_ISL_473623\|2020- | .................... | .................... | .................... | .................... | .................... | .................... | |
| D | 2958 | hCoV-19/Scotland/CVR2907/2020\|EPI_ISL_473624\|2020- | .................... | .................... | .................... | .................... | .................... | .................... | |
| D | 2959 | hCoV-19/Scotland/CVR2915/2020\|EPI_ISL_473628\|2020- | .................... | .................... | .................... | .................... | .................... | .................... | |
| D | 2960 | hCoV-19/Scotland/CVR2916/2020\|EPI_ISL_473629\|2020- | .................... | .................... | .................... | .................... | .................... | .................... | |
| D | 2961 | hCoV-19/Scotland/CVR2917/2020\|EPI_ISL_473630\|2020- | .................... | .A.................. | .................... | .................... | .................... | .................... | |
| D | 2962 | hCoV-19/Scotland/CVR2918/2020\|EPI_ISL_473631\|2020- | .................... | .................... | .................... | .................... | .................... | .................... | |
| D | 2963 | hCoV-19/Scotland/CVR2958/2020\|EPI_ISL_473632\|2020- | .................... | .................... | .................... | .................... | .................... | .................... | |
| D | 2964 | hCoV-19/Scotland/CVR2959/2020\|EPI_ISL_473633\|2020- | .................... | .A.................. | .................... | .................... | .................... | .................... | |
| D | 2965 | hCoV-19/Scotland/CVR2961/2020\|EPI_ISL_473635\|2020- | .................... | .................... | .................... | .................... | .................... | .................... | |
| D | 2966 | hCoV-19/Scotland/CVR2962/2020\|EPI_ISL_473636\|2020- | .................... | .................... | .................... | .................... | .................... | .................... | |
| D | 2967 | hCoV-19/Scotland/CVR2965/2020\|EPI_ISL_473637\|2020- | .................... | .................... | .................... | .................... | .................... | .................... | |
| D | 2968 | hCoV-19/Scotland/CVR2998/2020\|EPI_ISL_473642\|2020- | .................... | .................... | .................... | .................... | .................... | .................... | |
| D | 2969 | hCoV-19/Scotland/CVR3000/2020\|EPI_ISL_473643\|2020- | .................... | .................... | .................... | .................... | .................... | .................... | |
| D | 2970 | hCoV-19/Scotland/CVR3255/2020\|EPI_ISL_473645\|2020- | .................... | .................... | .................... | .................... | .................... | .................... | |
| D | 2971 | hCoV-19/Scotland/CVR3258/2020\|EPI_ISL_473646\|2020- | .................... | ...................A | .................... | .................... | .................... | .................... | 1 |
| D | 2972 | hCoV-19/Scotland/CVR3488/2020\|EPI_ISL_473647\|2020- | .................... | .A.................. | .................... | .................... | .................... | .................... | |
| D | 2976 | hCoV-19/Scotland/CVR3513/2020\|EPI_ISL_473653\|2020- | .................... | .................... | .................... | .................... | .................... | .................... | |
| D | 2977 | hCoV-19/Scotland/CVR3514/2020\|EPI_ISL_473654\|2020- | .................... | .................... | .................... | .................... | .................... | .................... | |
| D | 2978 | hCoV-19/Scotland/CVR3515/2020\|EPI_ISL_473655\|2020- | .................... | .................... | .................... | .................... | .................... | .................... | |
| D | 2979 | hCoV-19/Scotland/CVR3516/2020\|EPI_ISL_473656\|2020- | .................... | .................... | .................... | .................... | .................... | .................... | |
| D | 2980 | hCoV-19/Scotland/CVR3519/2020\|EPI_ISL_473659\|2020- | .................... | .................... | .................... | .................... | .................... | .................... | |
| D | 2981 | hCoV-19/Scotland/CVR3521/2020\|EPI_ISL_473660\|2020- | .................... | .................... | .................... | .................... | .................... | .................... | |
| D | 2982 | hCoV-19/Scotland/CVR3524/2020\|EPI_ISL_473662\|2020- | .................... | ..............C..... | .................... | .................... | .................... | .................... | |
| D | 2983 | hCoV-19/Scotland/CVR3525/2020\|EPI_ISL_473663\|2020- | .................... | .................... | .................... | .................... | .................... | .................... | |
| D | 2984 | hCoV-19/Scotland/CVR3528/2020\|EPI_ISL_473666\|2020- | .................... | .................... | .................... | .................... | .................... | .................... | |
| D | 2985 | hCoV-19/Scotland/CVR3529/2020\|EPI_ISL_473667\|2020- | .................... | .................... | .................... | .................... | .................... | .................... | |
| D | 2986 | hCoV-19/Scotland/CVR3532/2020\|EPI_ISL_473670\|2020- | .................... | .................... | .................... | .................... | .................... | .................... | |
| D | 2989 | hCoV-19/Scotland/CVR3537/2020\|EPI_ISL_473674\|2020- | .................... | .................... | .................... | .................... | .................... | .................... | |
| D | 2990 | hCoV-19/Scotland/CVR3539/2020\|EPI_ISL_473676\|2020- | .................... | .................... | .................... | .................... | .................... | .................... | |
| D | 2991 | hCoV-19/Scotland/CVR3544/2020\|EPI_ISL_473679\|2020- | .................... | .................... | .................... | .................... | .................... | .................... | |
| D | 2993 | hCoV-19/Scotland/CVR3546/2020\|EPI_ISL_473681\|2020- | .................... | .................... | .................... | .................... | .................... | .................... | |
| D | 2994 | hCoV-19/Scotland/CVR3559/2020\|EPI_ISL_473694\|2020- | .................... | .A.................. | .................... | .................... | .................... | .................... | |
| D | 2995 | hCoV-19/Scotland/CVR3562/2020\|EPI_ISL_473697\|2020- | .................... | .................... | .................... | .................... | .................... | .................... | |
| D | 3004 | hCoV-19/Spain/CM-IBV-003758/2020\|EPI_ISL_474933\|20 | .................... | .................... | .................... | .................... | .................... | .................... | |
| D | 3005 | hCoV-19/Spain/MC-IBV-002894/2020\|EPI_ISL_474934\|20 | .................... | .................... | .................... | .................... | .................... | .................... | |
| D | 3006 | hCoV-19/Spain/AN-IBV-003033/2020\|EPI_ISL_474935\|20 | .................... | .................... | .................... | .................... | .................... | .................... | |
| D | 3007 | hCoV-19/Spain/AN-IBV-002885/2020\|EPI_ISL_474936\|20 | .................... | .................... | .................... | .................... | .................... | .................... | |
| D | 3008 | hCoV-19/Spain/AN-IBV-002918/2020\|EPI_ISL_474939\|20 | .................... | .................... | .................... | .................... | .................... | .................... | |
| D | 3016 | hCoV-19/Spain/AN-IBV-003029/2020\|EPI_ISL_474950\|20 | .................... | .................... | .................... | .................... | .................... | .................... | |
| D | 3023 | hCoV-19/Israel/CVL-n-6744/2020\|EPI_ISL_474958\|2020 | .................... | .................... | .................... | .................... | .................... | .................... | |
| D | 3024 | hCoV-19/Israel/CVL-n-6750/2020\|EPI_ISL_474959\|2020 | .................... | .................... | .................... | .................... | .................... | .................... | |
| D | 3025 | hCoV-19/Israel/CVL-n-6751/2020\|EPI_ISL_474960\|2020 | .................... | .................... | .................... | .................... | .................... | .................... | |
| D | 3026 | hCoV-19/Israel/CVL-n-6051/2020\|EPI_ISL_474961\|2020 | .................... | .................... | .................... | .................... | .................... | .................... | |
| D | 3027 | hCoV-19/Israel/CVL-n-6656/2020\|EPI_ISL_474962\|2020 | .................... | .................... | .................... | .................... | .................... | .................... | |
| D | 3028 | hCoV-19/Israel/CVL-n-7216/2020\|EPI_ISL_474963\|2020 | .................... | .................... | .................... | .................... | .................... | .................... | |
| D | 3029 | hCoV-19/Israel/CVL-n-7175/2020\|EPI_ISL_474964\|2020 | .................... | .................... | .................... | .................... | .................... | .................... | |
| D | 3030 | hCoV-19/Israel/CVL-n-5939/2020\|EPI_ISL_474965\|2020 | .................... | .................... | .................... | .................... | .................... | .................... | |
| D | 3031 | hCoV-19/Israel/CVL-n-11470/2020\|EPI_ISL_474966\|202 | .................... | .................... | .................... | .................... | .................... | .................... | |
| D | 3032 | hCoV-19/Israel/CVL-n-11482/2020\|EPI_ISL_474967\|202 | .................... | .................... | .................... | .................... | .................... | .................... | |
| D | 3033 | hCoV-19/Israel/CVL-n-13881/2020\|EPI_ISL_474968\|202 | .................... | .................... | .................... | .................... | .................... | .................... | |
| D | 3034 | hCoV-19/Israel/CVL-n-6100/2020\|EPI_ISL_474969\|2020 | .................... | .................... | .................... | .................... | .................... | .................... | |
| D | 3035 | hCoV-19/Israel/CVL-n-9331/2020\|EPI_ISL_474970\|2020 | .................... | .................... | .................... | .................... | .................... | .................... | |
| D | 3036 | hCoV-19/Israel/CVL-n-9560/2020\|EPI_ISL_474971\|2020 | .................... | .................... | .................... | .................... | .................... | .................... | |
| D | 3037 | hCoV-19/Israel/CVL-n-11475/2020\|EPI_ISL_474972\|202 | .................... | .................... | .................... | .................... | .................... | .................... | |
| D | 3038 | hCoV-19/Israel/CVL-n-5853/2020\|EPI_ISL_474973\|2020 | .................... | .................... | .................... | .................... | .................... | .................... | |
| D | 3039 | hCoV-19/Israel/CVL-n-6035/2020\|EPI_ISL_474974\|2020 | .................... | .................... | .................... | .................... | .................... | .................... | |
| D | 3040 | hCoV-19/Israel/CVL-n-6108/2020\|EPI_ISL_474975\|2020 | .................... | .................... | .................... | .................... | .................... | .................... | |
| D | 3041 | hCoV-19/Israel/CVL-n-6735/2020\|EPI_ISL_474977\|2020 | .................... | .................... | .................... | .................... | .................... | .................... | |
| D | 3042 | hCoV-19/Israel/CVL-n-7154/2020\|EPI_ISL_474978\|2020 | .................... | .................... | .................... | .................... | .................... | .................... | |
| D | 3043 | hCoV-19/Israel/CVL-n-5826/2020\|EPI_ISL_474979\|2020 | .................... | .................... | .................... | .................... | .................... | .................... | |
| D | 3044 | hCoV-19/Israel/CVL-n-6005/2020\|EPI_ISL_474980\|2020 | .................... | .................... | .................... | .................... | .................... | .................... | |
| D | 3045 | hCoV-19/Israel/CVL-n-6140/2020\|EPI_ISL_474981\|2020 | .................... | .................... | .................... | .................... | .................... | .................... | |
| D | 3046 | hCoV-19/Israel/CVL-n-8405/2020\|EPI_ISL_474982\|2020 | .................... | .................... | .................... | .................... | .................... | .................... | |
| D | 3047 | hCoV-19/Israel/CVL-n-8414/2020\|EPI_ISL_474983\|2020 | .................... | .................... | .................... | .................... | .................... | .................... | |
| D | 3048 | hCoV-19/Israel/CVL-n-10864/2020\|EPI_ISL_474984\|202 | .................... | .................... | .................... | .................... | .................... | .................... | |

| | | | | | | | | |
|---|---|---|---|---|---|---|---|---|
| D | 3049 hCoV-19/Israel/CVL-n-11468/2020\|EPI_ISL_474985\|202 | ········T··········· | ···················· | ···················· | ···················· | ···················· | ···················· | |
| D | 3050 hCoV-19/Israel/CVL-n-5856/2020\|EPI_ISL_474986\|2020 | ···················· | ···················· | ···················· | ···················· | ···················· | ···················· | |
| D | 3051 hCoV-19/Israel/CVL-n-6102/2020\|EPI_ISL_474987\|2020 | ···················· | ···················· | ···················· | ···················· | ···················· | ···················· | |
| D | 3052 hCoV-19/Israel/CVL-n-6118/2020\|EPI_ISL_474988\|2020 | ···················· | ···················· | ···················· | ···················· | ···················· | ···················· | |
| D | 3053 hCoV-19/Israel/CVL-n-7224/2020\|EPI_ISL_474989\|2020 | ···················· | ···················· | ···················· | ···················· | ···················· | ···················· | |
| D | 3054 hCoV-19/Israel/CVL-n-9434/2020\|EPI_ISL_474990\|2020 | ···················· | ···················· | ···················· | ···················· | ···················· | ···················· | |
| D | 3055 hCoV-19/Israel/CVL-n-10261/2020\|EPI_ISL_474991\|202 | ···················· | ···················· | ···················· | ···················· | ···················· | ···················· | |
| D | 3056 hCoV-19/Israel/CVL-n-6106/2020\|EPI_ISL_474992\|2020 | ···················· | ···················· | ···················· | ···················· | ···················· | ···················· | |
| D | 3057 hCoV-19/Israel/CVL-n-6126/2020\|EPI_ISL_474993\|2020 | ···················· | ···················· | ···················· | ···················· | ···················· | ···················· | |
| D | 3058 hCoV-19/Israel/CVL-n-6131/2020\|EPI_ISL_474994\|2020 | ···················· | ···················· | ···················· | ···················· | ···················· | ···················· | |
| D | 3059 hCoV-19/Israel/CVL-n-14266/2020\|EPI_ISL_474995\|202 | ···················· | ···················· | ···················· | ···················· | ···················· | ···················· | |
| D | 3060 hCoV-19/Israel/CVL-n-7169/2020\|EPI_ISL_474996\|2020 | ···················· | ···················· | ···················· | ···················· | ···················· | ···················· | |
| D | 3061 hCoV-19/Israel/CVL-n-7467/2020\|EPI_ISL_474997\|2020 | ···················· | ···················· | ···················· | ···················· | ···················· | ···················· | |
| D | 3062 hCoV-19/Israel/CVL-n-7905/2020\|EPI_ISL_474998\|2020 | ···················· | ···················· | ···················· | ···················· | ···················· | ···················· | |
| D | 3063 hCoV-19/Israel/CVL-n-6104/2020\|EPI_ISL_474999\|2020 | ···················· | ···················· | ···················· | ···················· | ···················· | ···················· | |
| D | 3064 hCoV-19/Singapore/470/2020\|EPI_ISL_477170\|2020-02- | ···················· | ···················· | ···················· | ···················· | ···················· | ···················· | |
| D | 3065 hCoV-19/Singapore/481/2020\|EPI_ISL_477171\|2020-03- | ···················· | ···················· | ···················· | ···················· | ···················· | ···················· | |
| D | 3066 hCoV-19/Singapore/431nan/2020\|EPI_ISL_477172\|2020- | ···················· | ···················· | ···················· | ···················· | ···················· | ···················· | |
| D | 3067 hCoV-19/Singapore/451nan/2020\|EPI_ISL_477174\|2020- | ···················· | ···················· | ···················· | ···················· | ···················· | ···················· | |
| D | 3068 hCoV-19/Singapore/485/2020\|EPI_ISL_477175\|2020-03- | ···················· | ···················· | ···················· | ···················· | ···················· | ···················· | |
| D | 3069 hCoV-19/Singapore/159nan/2020\|EPI_ISL_477177\|2020- | ···················· | ···················· | ···················· | ···················· | ···················· | ···················· | |
| D | 3070 hCoV-19/Singapore/278nan/2020\|EPI_ISL_477178\|2020- | ···················· | ···················· | ···················· | ···················· | ···················· | ···················· | |
| D | 3071 hCoV-19/India/GJ-GBRC234/2020\|EPI_ISL_477183\|2020- | ···················· | ···················· | ···················· | ···················· | ···················· | ···················· | |
| D | 3072 hCoV-19/Singapore/489/2020\|EPI_ISL_477184\|2020-04- | ···················· | ···················· | ···················· | ···················· | ···················· | ···················· | |
| D | 3073 hCoV-19/USA/MN-MDH-1065/2020\|EPI_ISL_462910\|2020-0 | ···················· | ·A·················· | ···················· | ···················· | ···················· | ···················· | |
| D | 3074 hCoV-19/USA/MN-MDH-1066/2020\|EPI_ISL_462911\|2020-0 | ···················· | ·A·················· | ···················· | ···················· | ···················· | ···················· | |
| D | 3075 hCoV-19/USA/CO-WYPHL-00036/2020\|EPI_ISL_462912\|202 | ···················· | ···················· | ···················· | ···················· | ···················· | ···················· | |
| D | 3076 hCoV-19/USA/WY-WYPHL-00022/2020\|EPI_ISL_462915\|202 | ···················· | ···················· | ···················· | ···················· | ···················· | ···················· | |
| D | 3077 hCoV-19/USA/WY-WYPHL-00024/2020\|EPI_ISL_462917\|202 | ···················· | ···················· | ···················· | ···················· | ···················· | ···················· | |
| D | 3078 hCoV-19/USA/WY-WYPHL-00025/2020\|EPI_ISL_462918\|202 | ···················· | ···················· | ···················· | ···················· | ···················· | ···················· | |
| D | 3079 hCoV-19/USA/WY-WYPHL-00026/2020\|EPI_ISL_462919\|202 | ···················· | ···················· | ···················· | ···················· | ···················· | ···················· | |
| D | 3080 hCoV-19/USA/WY-WYPHL-00031/2020\|EPI_ISL_462924\|202 | ···················· | ···················· | ···················· | ···················· | ···················· | ···················· | |
| D | 3081 hCoV-19/USA/WY-WYPHL-00034/2020\|EPI_ISL_462927\|202 | ···················· | ···················· | ···················· | ···················· | ···················· | ···················· | |
| D | 3082 hCoV-19/USA/WY-WYPHL-00037/2020\|EPI_ISL_462929\|202 | ···················· | ···················· | ···················· | ···················· | ···················· | ···················· | |
| D | 3083 hCoV-19/USA/WY-WYPHL-00040/2020\|EPI_ISL_462932\|202 | ···················· | ···················· | ···················· | ···················· | ···················· | ···················· | |
| D | 3084 hCoV-19/USA/WY-WYPHL-00041/2020\|EPI_ISL_462933\|202 | ···················· | ···················· | ···················· | ···················· | ···················· | ···················· | |
| D | 3085 hCoV-19/USA/WY-WYPHL-00043/2020\|EPI_ISL_462935\|202 | ···················· | ···················· | ···················· | ···················· | ···················· | ···················· | |
| D | 3086 hCoV-19/USA/WY-WYPHL-00044/2020\|EPI_ISL_462936\|202 | ···················· | ···················· | ···················· | ···················· | ···················· | ···················· | |
| D | 3087 hCoV-19/USA/WY-WYPHL-00048/2020\|EPI_ISL_462940\|202 | ···················· | ···················· | ···················· | ···················· | ···················· | ···················· | |
| D | 3088 hCoV-19/Canada/ON-UHTC_0061/2020\|EPI_ISL_464001\|20 | ···················· | ···················· | ···················· | ···················· | ···················· | ···················· | |
| D | 3089 hCoV-19/Canada/ON-UHTC_0053/2020\|EPI_ISL_464004\|20 | ···················· | ···················· | ···················· | ···················· | ···················· | ···················· | |
| D | 3090 hCoV-19/Canada/ON-UHTC_0063/2020\|EPI_ISL_464007\|20 | ···················· | ···················· | ···················· | ···················· | ···················· | ···················· | |
| D | 3091 hCoV-19/Canada/ON-UHTC_0068/2020\|EPI_ISL_464008\|20 | ···················· | ···················· | ···················· | ···················· | ···················· | ···················· | |
| D | 3092 hCoV-19/Canada/ON-UHTC_0091/2020\|EPI_ISL_464011\|20 | ···················· | ···················· | ···················· | ···················· | ···················· | ···················· | |
| D | 3093 hCoV-19/Canada/ON-UHTC_0047/2020\|EPI_ISL_464012\|20 | ···················· | ···················· | ···················· | ···················· | ···················· | ···················· | |
| D | 3094 hCoV-19/Canada/ON-UHTC_0049/2020\|EPI_ISL_464013\|20 | ···················· | ···················· | ···················· | ···················· | ···················· | ···················· | |
| D | 3095 hCoV-19/Canada/ON-UHTC_0041/2020\|EPI_ISL_464037\|20 | ···················· | ···················· | ···················· | ···················· | ···················· | ···················· | |
| D | 3096 hCoV-19/Canada/ON-UHTC_0066/2020\|EPI_ISL_464038\|20 | ···················· | ···················· | ···················· | ···················· | ···················· | ···················· | |
| D | 3097 hCoV-19/Canada/ON-UHTC_0080/2020\|EPI_ISL_464040\|20 | ···················· | ···················· | ···················· | ···················· | ···················· | ···················· | |
| D | 3098 hCoV-19/Canada/ON-UHTC_0042/2020\|EPI_ISL_464041\|20 | ···················· | ···················· | ···················· | ···················· | ···················· | ···················· | |
| D | 3099 hCoV-19/Canada/ON-UHTC_0057/2020\|EPI_ISL_464042\|20 | ···················· | ···················· | ···················· | ···················· | ···················· | ···················· | |
| D | 3100 hCoV-19/Canada/ON-UHTC_0055/2020\|EPI_ISL_464046\|20 | ···················· | ···················· | ···················· | ···················· | ···················· | ···················· | |
| D | 3101 hCoV-19/Canada/ON-UHTC_0036/2020\|EPI_ISL_464047\|20 | ···················· | ···················· | ···················· | ···················· | ···················· | ···················· | |
| D | 3102 hCoV-19/Canada/ON-UHTC_0062/2020\|EPI_ISL_464048\|20 | ···················· | ···················· | ···················· | ···················· | ···················· | ···················· | |
| D | 3103 hCoV-19/Canada/ON-UHTC_0054/2020\|EPI_ISL_464050\|20 | ···················· | ···················· | ···················· | ···················· | ···················· | ···················· | |
| D | 3104 hCoV-19/Canada/ON-UHTC_0088/2020\|EPI_ISL_464054\|20 | ···················· | ···················· | ···················· | ···················· | ···················· | ···················· | |
| D | 3105 hCoV-19/Canada/ON-UHTC_0052/2020\|EPI_ISL_464056\|20 | ···················· | ···················· | ···················· | ···················· | ···················· | ···················· | |
| D | 3106 hCoV-19/Canada/ON-UHTC_0086/2020\|EPI_ISL_464058\|20 | ···················· | ···················· | ···················· | ···················· | ···················· | ···················· | |
| D | 3107 hCoV-19/England/20120013604/2020\|EPI_ISL_465155\|20 | ···················· | ···················· | ···················· | ···················· | ···················· | ···················· | |
| D | 3108 hCoV-19/Bangladesh/BCSIR-NILMRC-053/2020\|EPI_ISL_4 | ···················· | ···················· | ···················· | ···················· | ···················· | ···················· | |
| D | 3109 hCoV-19/Bangladesh/BCSIR-NILMRC-059/2020\|EPI_ISL_4 | ···················· | ···················· | ···················· | ···················· | ···················· | ···················· | |
| D | 3110 hCoV-19/England/20169001704/2020\|EPI_ISL_465167\|20 | ···················· | ···················· | ···················· | ···················· | ···················· | ···················· | |
| D | 3111 hCoV-19/England/20169001804/2020\|EPI_ISL_465168\|20 | ···················· | ···················· | ···················· | ···················· | ···················· | ···················· | |
| D | 3112 hCoV-19/England/20169002104/2020\|EPI_ISL_465169\|20 | ···················· | ···················· | ···················· | ···················· | ···················· | ···················· | |
| D | 3116 hCoV-19/England/20169014104/2020\|EPI_ISL_465195\|20 | ···················· | ···················· | ···················· | ···················· | ······N··NN········· | ···················· | 1 |
| D | 3117 hCoV-19/England/20169014604/2020\|EPI_ISL_465196\|20 | ···················· | ···················· | ···················· | ···················· | ···················· | ···················· | |
| D | 3120 hCoV-19/England/20169015304/2020\|EPI_ISL_465199\|20 | ···················· | ···················· | ···················· | ···················· | ···················· | ···················· | |
| D | 3121 hCoV-19/England/20170048604/2020\|EPI_ISL_465302\|20 | ···················· | ···················· | ···················· | ···················· | ···················· | ···················· | |
| D | 3124 hCoV-19/England/20172000204/2020\|EPI_ISL_465309\|20 | ···················· | ···················· | ···················· | ···················· | ···················· | ···················· | |
| D | 3125 hCoV-19/England/20172000504/2020\|EPI_ISL_465311\|20 | ···················· | ···················· | ···················· | ···················· | ···················· | ···················· | |
| D | 3126 hCoV-19/England/20172000604/2020\|EPI_ISL_465312\|20 | ···················· | ···················· | ···················· | ···················· | ···················· | ···················· | |
| D | 3127 hCoV-19/England/20172000704/2020\|EPI_ISL_465313\|20 | ···················· | ···················· | ···················· | ···················· | ···················· | ···················· | |
| D | 3128 hCoV-19/England/20172001004/2020\|EPI_ISL_465315\|20 | ···················· | ···················· | ···················· | ···················· | ···················· | ···················· | |
| D | 3129 hCoV-19/England/20172002104/2020\|EPI_ISL_465318\|20 | ···················· | ···················· | ···················· | ···················· | ···················· | ···················· | |
| D | 3130 hCoV-19/England/20172002204/2020\|EPI_ISL_465319\|20 | ···················· | ···················· | ···················· | ···················· | ···················· | ···················· | |
| D | 3132 hCoV-19/England/20172002704/2020\|EPI_ISL_465322\|20 | ···················· | ···················· | ···················· | ···················· | ···················· | ···················· | |
| D | 3133 hCoV-19/England/20172003004/2020\|EPI_ISL_465324\|20 | ···················· | ···················· | ···················· | ···················· | ·NN················· | ·N·················· | |
| D | 3134 hCoV-19/England/20172009004/2020\|EPI_ISL_465332\|20 | ···················· | ···················· | ···················· | ···················· | ···················· | ···················· | |

| | | | | | | | | |
|---|---|---|---|---|---|---|---|---|
| D | 3136 | hCoV-19/England/20172042904/2020\|EPI_ISL_465343\|20 | ···················· | ···················· | ····················· | ···················· | ···················· | ···················· |
| D | 3137 | hCoV-19/England/20172043604/2020\|EPI_ISL_465344\|20 | ···················· | ···················· | ····················· | ···················· | ···················· | ···················· |
| D | 3138 | hCoV-19/England/20172055404/2020\|EPI_ISL_465361\|20 | ···················· | ···················· | ····················· | ···················· | ···················· | ···················· |
| D | 3139 | hCoV-19/England/20172059204/2020\|EPI_ISL_465370\|20 | ···················· | ···················· | ····················· | ···················· | ···················· | NN·················· |
| D | 3140 | hCoV-19/England/20172060104/2020\|EPI_ISL_465371\|20 | ···················· | ···················· | ····················· | ···················· | ···················· | ···················· |
| D | 3141 | hCoV-19/England/20174003604/2020\|EPI_ISL_465381\|20 | ···················· | ···················· | ····················· | ···················· | ···················· | ···················· |
| D | 3142 | hCoV-19/England/20174003904/2020\|EPI_ISL_465382\|20 | ···················· | ···················· | ····················· | ···················· | ···················· | ···················· |
| D | 3143 | hCoV-19/England/20174004204/2020\|EPI_ISL_465383\|20 | ···················· | ···················· | ····················· | ···················· | ···················· | ···················· |
| D | 3145 | hCoV-19/England/20174004504/2020\|EPI_ISL_465386\|20 | ···················· | ···················· | ····················· | ···················· | ···················· | ···················· |
| D | 3147 | hCoV-19/England/20174019404/2020\|EPI_ISL_465389\|20 | ···················· | ···················· | ····················· | ···················· | ···················· | ···················· |
| D | 3148 | hCoV-19/England/20164082704/2020\|EPI_ISL_466485\|20 | ···················· | ···················· | ····················· | ···················· | ···················· | ···················· |
| D | 3149 | hCoV-19/England/20166000104/2020\|EPI_ISL_466492\|20 | ···················· | ···················· | ····················· | ···················· | ···················· | ···················· |
| D | 3150 | hCoV-19/England/20166000204/2020\|EPI_ISL_466493\|20 | ···················· | ···················· | ····················· | ···················· | ···················· | ···················· |
| D | 3151 | hCoV-19/England/20166000704/2020\|EPI_ISL_466496\|20 | ···················· | ···················· | ····················· | ···················· | ···················· | ···················· |
| D | 3152 | hCoV-19/England/20166000904/2020\|EPI_ISL_466498\|20 | ···················· | ···················· | ····················· | ···················· | ···················· | ···················· |
| D | 3153 | hCoV-19/Bangladesh/BCSIR-NILMRC-058/2020\|EPI_ISL_4 | ···················· | ···················· | ····················· | ···················· | ···················· | ···················· |
| D | 3154 | hCoV-19/Bangladesh/BCSIR-NILMRC-067/2020\|EPI_ISL_4 | ···················· | ·A·················· | ····················· | ···················· | ···················· | ··A················· |
| D | 3155 | hCoV-19/Bangladesh/BCSIR-NILMRC-069/2020\|EPI_ISL_4 | ···················· | ·A·················· | ····················· | ···················· | ···················· | ··A················· |
| D | 3156 | hCoV-19/Bangladesh/BCSIR-NILMRC-070/2020\|EPI_ISL_4 | ···················· | ·A·················· | ····················· | ···················· | ···················· | ··A················· |
| D | 3157 | hCoV-19/Bangladesh/BCSIR-NILMRC-076/2020\|EPI_ISL_4 | ···················· | ···················· | ····················· | ···················· | ···················· | ···················· |
| D | 3158 | hCoV-19/Bangladesh/BCSIR-NILMRC-077/2020\|EPI_ISL_4 | ···················· | ···················· | ····················· | ···················· | ···················· | ···················· |
| D | 3159 | hCoV-19/USA/CA-IGI-0045/2020\|EPI_ISL_466648\|2020-0 | ···················· | ···················· | ····················· | ···················· | ···················· | ···················· |
| D | 3160 | hCoV-19/Bangladesh/BCSIR-NILMRC-082/2020\|EPI_ISL_4 | ···················· | ···················· | ····················· | ···················· | ···················· | ···················· |
| D | 3161 | hCoV-19/Bangladesh/BCSIR-NILMRC-085/2020\|EPI_ISL_4 | ···················· | ···················· | ····················· | ···················· | ···················· | ···················· |
| D | 3162 | hCoV-19/USA/NE-10744/2020\|EPI_ISL_466652\|2020-05-0 | ···················· | ···················· | ····················· | ···················· | ···················· | ···················· |
| D | 3163 | hCoV-19/USA/NE-10746/2020\|EPI_ISL_466653\|2020-05-0 | ···················· | ···················· | ····················· | ···················· | ···················· | ···················· |
| D | 3164 | hCoV-19/USA/NE-10940/2020\|EPI_ISL_466655\|2020-05-0 | ···················· | ···················· | ····················· | ···················· | ···················· | ···················· |
| D | 3166 | hCoV-19/USA/NE-11323/2020\|EPI_ISL_466661\|2020-05-0 | ···················· | ···················· | ····················· | ···················· | ···················· | ···················· |
| D | 3168 | hCoV-19/USA/NE-10882/2020\|EPI_ISL_466666\|2020-05-0 | ···················· | ···················· | ····················· | ···················· | ···················· | ···················· |
| D | 3169 | hCoV-19/USA/NE-10900/2020\|EPI_ISL_466667\|2020-05-0 | ···················· | ···················· | ····················· | ···················· | ···················· | ···················· |
| D | 3170 | hCoV-19/USA/NE-10928/2020\|EPI_ISL_466668\|2020-05-0 | ···················· | ···················· | ····················· | ···················· | ···················· | ···················· |
| D | 3171 | hCoV-19/USA/NE-10933/2020\|EPI_ISL_466669\|2020-05-0 | ···················· | ···················· | ····················· | ···················· | ···················· | ···················· |
| D | 3172 | hCoV-19/USA/NE-10939/2020\|EPI_ISL_466670\|2020-05-0 | ···················· | ···················· | ····················· | ···················· | ···················· | ···················· |
| D | 3175 | hCoV-19/USA/NE-11308/2020\|EPI_ISL_466675\|2020-05-0 | ···················· | ···················· | ····················· | ···················· | ···················· | ···················· |
| D | 3176 | hCoV-19/Bangladesh/BCSIR-NILMRC-075/2020\|EPI_ISL_4 | ···················· | ···················· | ····················· | ···················· | ···················· | ···················· |
| D | 3177 | hCoV-19/Bangladesh/BCSIR-NILMRC-078/2020\|EPI_ISL_4 | ···················· | ···················· | ····················· | ···················· | ···················· | ···················· |
| D | 3178 | hCoV-19/Bangladesh/BCSIR-NILMRC-081/2020\|EPI_ISL_4 | ···················· | ···················· | ····················· | ···················· | ···················· | ···················· |
| D | 3179 | hCoV-19/Canada/BC_00696199/2020\|EPI_ISL_466699\|202 | ···················· | ···················· | ····················· | ···················· | ···················· | ···················· |
| D | 3180 | hCoV-19/USA/CA-SR0436/2020\|EPI_ISL_467964\|2020-03- | ···················· | ···················· | ····················· | ···················· | ···················· | ···················· |
| D | 3181 | hCoV-19/USA/CA-SR0438/2020\|EPI_ISL_467965\|2020-03- | ···················· | ···················· | ····················· | ···················· | ···················· | ···················· |
| D | 3182 | hCoV-19/USA/CA-SR0439/2020\|EPI_ISL_467966\|2020-03- | ···················· | ···················· | ····················· | ···················· | ···················· | ···················· |
| D | 3183 | hCoV-19/USA/CA-SR0440/2020\|EPI_ISL_467967\|2020-03- | ···················· | ···················· | ····················· | ···················· | ···················· | ···················· |
| D | 3184 | hCoV-19/USA/CA-SR0443/2020\|EPI_ISL_467968\|2020-03- | ···················· | ···················· | ····················· | ···················· | ···················· | ···················· |
| D | 3185 | hCoV-19/USA/CA-SR0444/2020\|EPI_ISL_467969\|2020-03- | ···················· | ···················· | ····················· | ···················· | ···················· | ···················· |
| D | 3186 | hCoV-19/USA/CA-SR0445/2020\|EPI_ISL_467970\|2020-03- | ···················· | ···················· | ····················· | ···················· | ···················· | ···················· |
| D | 3187 | hCoV-19/USA/CA-SR0447/2020\|EPI_ISL_467971\|2020-03- | ···················· | ···················· | ····················· | ···················· | ···················· | ···················· |
| D | 3188 | hCoV-19/USA/CA-SR0448/2020\|EPI_ISL_467972\|2020-04- | ···················· | ···················· | ····················· | ···················· | ···················· | ···················· |
| D | 3189 | hCoV-19/USA/CA-SR0449/2020\|EPI_ISL_467973\|2020-03- | ···················· | ···················· | ····················· | ···················· | ···················· | ···················· |
| D | 3190 | hCoV-19/USA/CA-SR0450/2020\|EPI_ISL_467974\|2020-03- | ···················· | ···················· | ····················· | ···················· | ···················· | ···················· |
| D | 3191 | hCoV-19/USA/CA-SR0452/2020\|EPI_ISL_467975\|2020-03- | ···················· | ···················· | ····················· | ···················· | ···················· | ···················· |
| D | 3192 | hCoV-19/USA/CA-SR0453/2020\|EPI_ISL_467976\|2020-05- | ···················· | ···················· | ····················· | ···················· | ···················· | ···················· |
| D | 3193 | hCoV-19/USA/CA-SR0455/2020\|EPI_ISL_467978\|2020-03- | ···················· | ···················· | ····················· | ···················· | ···················· | ···················· |
| D | 3194 | hCoV-19/USA/CA-SR0457/2020\|EPI_ISL_467979\|2020-03- | ···················· | ···················· | ····················· | ···················· | ···················· | ···················· |
| D | 3195 | hCoV-19/USA/CA-SR0458/2020\|EPI_ISL_467980\|2020-04- | ···················· | ···················· | ····················· | ···················· | ···················· | ···················· |
| D | 3196 | hCoV-19/USA/CA-SR0459/2020\|EPI_ISL_467981\|2020-05- | ···················· | ·A·················· | ····················· | ···················· | ···················· | ···················· |
| D | 3197 | hCoV-19/USA/CA-SR0460/2020\|EPI_ISL_467982\|2020-03- | ···················· | ···················· | ····················· | ···················· | ···················· | ···················· |
| D | 3198 | hCoV-19/USA/CA-SR0461/2020\|EPI_ISL_467983\|2020-03- | ···················· | ···················· | ····················· | ···················· | ···················· | ···················· |
| D | 3199 | hCoV-19/USA/CA-SR0463/2020\|EPI_ISL_467984\|2020-05- | ···················· | ·A·················· | ····················· | ···················· | ···················· | ···················· |
| D | 3200 | hCoV-19/Australia/SAP015/2020\|EPI_ISL_467990\|2020- | ···················· | ···················· | ····················· | ···················· | ···················· | ···················· |
| D | 3201 | hCoV-19/Australia/SAP272/2020\|EPI_ISL_467991\|2020- | ···················· | ···················· | ····················· | ···················· | ···················· | ···················· |
| D | 3202 | hCoV-19/Australia/SAP018/2020\|EPI_ISL_467994\|2020- | ···················· | ···················· | ····················· | ···················· | ···················· | ···················· |
| D | 3203 | hCoV-19/Australia/SAP220/2020\|EPI_ISL_467995\|2020- | ···················· | ···················· | ····················· | ···················· | ···················· | ···················· |
| D | 3204 | hCoV-19/Australia/SAP276/2020\|EPI_ISL_467996\|2020- | ···················· | ···················· | ····················· | ···················· | ···················· | ···················· |
| D | 3205 | hCoV-19/Australia/SAP245/2020\|EPI_ISL_467999\|2020- | ···················· | ···················· | ····················· | ···················· | ···················· | ···················· |
| D | 3206 | hCoV-19/Spain/CN-IBV-002593/2020\|EPI_ISL_474910\|20 | ···················· | ···················· | ····················· | ···················· | ···················· | ···················· |
| D | 3211 | hCoV-19/Spain/CN-IBV-002599/2020\|EPI_ISL_474915\|20 | ···················· | ···················· | ····················· | ···················· | ···················· | ···················· |
| D | 3222 | hCoV-19/USA/MO-WUSTL070/2020\|EPI_ISL_476018\|2020-0 | ···················· | ···················· | ····················· | ···················· | ···················· | ···················· |
| D | 3223 | hCoV-19/USA/MO-WUSTL066/2020\|EPI_ISL_476019\|2020-0 | ···················· | ···················· | ····················· | ···················· | ···················· | ···················· |
| D | 3224 | hCoV-19/USA/MO-WUSTL065/2020\|EPI_ISL_476020\|2020-0 | ···················· | ···················· | ····················· | ···················· | ···················· | ···················· |
| D | 3225 | hCoV-19/USA/MO-WUSTL032/2020\|EPI_ISL_476021\|2020-0 | ···················· | ···················· | ····················· | ···················· | ···················· | ···················· |
| D | 3226 | hCoV-19/India/MP-DRDE08/2020\|EPI_ISL_476022\|2020-0 | ···················· | ···················· | ····················· | ···················· | ···················· | ···················· |
| D | 3227 | hCoV-19/India/MP-DRDE115/2020\|EPI_ISL_476023\|2020- | ···················· | ···················· | ····················· | ···················· | ···················· | ···················· |
| D | 3228 | hCoV-19/USA/MI-MDHHS-SC20890/2020\|EPI_ISL_476027\|2 | ···················· | ···················· | ····················· | ···················· | ···················· | ···················· |
| D | 3229 | hCoV-19/USA/MI-MDHHS-SC20891/2020\|EPI_ISL_476028\|2 | ···················· | ··C················· | ····················· | ···················· | ···················· | ···················· |
| D | 3230 | hCoV-19/USA/MI-MDHHS-SC20892/2020\|EPI_ISL_476029\|2 | ···················· | ···················· | ····················· | ···················· | ···················· | ···················· |
| D | 3231 | hCoV-19/USA/MI-MDHHS-SC20893/2020\|EPI_ISL_476030\|2 | ···················· | ···················· | ····················· | ···················· | ···················· | ···················· |
| D | 3232 | hCoV-19/USA/MI-MDHHS-SC20895/2020\|EPI_ISL_476032\|2 | ···················· | ···················· | ····················· | ···················· | ···················· | ···················· |
| D | 3233 | hCoV-19/USA/MI-MDHHS-SC20896/2020\|EPI_ISL_476033\|2 | ···················· | ···················· | ····················· | ···················· | ···················· | ···················· |

| | | | | | | | | | |
|---|---|---|---|---|---|---|---|---|---|
| D | 3234 | hCoV-19/USA/MI-MDHHS-SC20897/2020\|EPI_ISL_476034\|2 | ···················· | ···················· | ···················· | ···················· | ···················· | ···················· | |
| D | 3235 | hCoV-19/Bangladesh/CHRF-0014/2020\|EPI_ISL_477127\|2 | ···················· | ···················· | ···················· | ···················· | ···················· | ···················· | |
| D | 3236 | hCoV-19/Bangladesh/CHRF-0015/2020\|EPI_ISL_477128\|2 | ···················· | ···················· | ···················· | ···················· | ···················· | ···················· | |
| D | 3237 | hCoV-19/Bangladesh/CHRF-0016/2020\|EPI_ISL_477129\|2 | ···················· | ···················· | ···················· | ···················· | ···················· | ···················· | |
| D | 3238 | hCoV-19/Bangladesh/CHRF-0017/2020\|EPI_ISL_477130\|2 | ···················· | ···················· | ···················· | ···················· | ···················· | ···················· | |
| D | 3239 | hCoV-19/Bangladesh/CHRF-0020/2020\|EPI_ISL_477131\|2 | ···················· | ···················· | ···················· | ···················· | ···················· | ···················· | |
| D | 3240 | hCoV-19/Bangladesh/CHRF-0022/2020\|EPI_ISL_477132\|2 | ···················· | ···················· | ···················· | ···················· | ···················· | ···················· | |
| D | 3241 | hCoV-19/Bangladesh/CHRF-0023/2020\|EPI_ISL_477133\|2 | ···················· | ···················· | ···················· | ···················· | ···················· | ···················· | |
| D | 3242 | hCoV-19/Bangladesh/CHRF-0024/2020\|EPI_ISL_477134\|2 | ···················· | ·A·················· | ···················· | ···················· | ···················· | ··A················· | |
| D | 3243 | hCoV-19/Bangladesh/CHRF-0025/2020\|EPI_ISL_477135\|2 | ···················· | ···················· | ···················· | ···················· | ···················· | ···················· | |
| D | 3244 | hCoV-19/Bangladesh/CHRF-0027/2020\|EPI_ISL_477136\|2 | ···················· | ···················· | ···················· | ···················· | ···················· | ···················· | |
| D | 3245 | hCoV-19/Bangladesh/CHRF-0029/2020\|EPI_ISL_477138\|2 | ···················· | ···················· | ···················· | ···················· | ···················· | ···················· | |
| D | 3246 | hCoV-19/Bangladesh/CHRF-0030/2020\|EPI_ISL_477139\|2 | ···················· | ·A·················· | ···················· | ···················· | ···················· | ··A················· | |
| D | 3247 | hCoV-19/Bangladesh/CHRF-0031/2020\|EPI_ISL_477140\|2 | ···················· | ···················· | ···················· | ···················· | ···················· | ···················· | |
| D | 3248 | hCoV-19/Senegal/68303/2020\|EPI_ISL_477143\|2020-06- | ···················· | ···················· | ···················· | ···················· | ···················· | ···················· | |
| D | 3249 | hCoV-19/Senegal/55728/2020\|EPI_ISL_477150\|2020-06- | ···················· | ···················· | ···················· | ···················· | ···················· | ···················· | |
| D | 3250 | hCoV-19/Senegal/64729/2020\|EPI_ISL_477155\|2020-06- | ···················· | ···················· | ···················· | ···················· | ···················· | ···················· | |
| D | 3251 | hCoV-19/Senegal/64916/2020\|EPI_ISL_477156\|2020-06- | ···················· | ···················· | ···················· | ···················· | ···················· | ···················· | |
| D | 3252 | hCoV-19/env/USA/FL-UF-20/2020\|EPI_ISL_477160\|2020- | ···················· | ···················· | ···················· | ···················· | ···················· | ···················· | |
| D | 3253 | hCoV-19/Egypt/CUNCI-HGC6I029/2020\|EPI_ISL_477161\|2 | ···················· | ···················· | ···················· | ···················· | ···················· | ···················· | |
| D | 3254 | hCoV-19/USA/FL-UF-19/2020\|EPI_ISL_477163\|2020-04-1 | ···················· | ···················· | ···················· | ···················· | ···················· | ···················· | |
| D | 3255 | hCoV-19/Georgia/Tb-7851/2020\|EPI_ISL_477169\|2020-0 | ···················· | ···················· | ···················· | ···················· | ···················· | ···················· | |
| D | 3256 | hCoV-19/Singapore/268nan/2020\|EPI_ISL_477180\|2020- | ···················· | ···················· | ···················· | ···················· | ···················· | ···················· | |
| D | 3257 | hCoV-19/Singapore/488/2020\|EPI_ISL_477182\|2020-04- | ···················· | ···················· | ···················· | ···················· | ···················· | ···················· | |
| D | 3258 | hCoV-19/Singapore/490/2020\|EPI_ISL_477187\|2020-04- | ···················· | ···················· | ···················· | ···················· | ···················· | ···················· | |
| D | 3259 | hCoV-19/Singapore/491/2020\|EPI_ISL_477188\|2020-04- | ···················· | ···················· | ···················· | ···················· | ···················· | ···················· | |
| D | 3260 | hCoV-19/Singapore/492/2020\|EPI_ISL_477189\|2020-04- | ···················· | ···················· | ···················· | ···················· | ···················· | ···················· | |
| D | 3261 | hCoV-19/Singapore/493/2020\|EPI_ISL_477190\|2020-04- | ···················· | ···················· | ···················· | ···················· | ···················· | ···················· | |
| D | 3262 | hCoV-19/Singapore/494/2020\|EPI_ISL_477191\|2020-04- | ···················· | ···················· | ···················· | ···················· | ···················· | ···················· | |
| D | 3263 | hCoV-19/Singapore/495/2020\|EPI_ISL_477192\|2020-04- | ···················· | ···················· | ···················· | ···················· | ···················· | ···················· | |
| D | 3264 | hCoV-19/Italy/APU-UniMI-45/2020\|EPI_ISL_477193\|202 | ···················· | ···················· | ···················· | ···················· | ···················· | ···················· | |
| D | 3265 | hCoV-19/Italy/BAS-UniMI-2167/2020\|EPI_ISL_477199\|2 | ···················· | ···················· | ···················· | ···················· | ···················· | ···················· | |
| D | 3266 | hCoV-19/USA/MN-MDH-1211/2020\|EPI_ISL_477300\|2020-0 | ···················· | ···················· | ···················· | ···················· | ···················· | ···················· | |
| D | 3267 | hCoV-19/USA/MN-MDH-1212/2020\|EPI_ISL_477301\|2020-0 | ···················· | ···················· | ···················· | ···················· | ···················· | ···················· | |
| D | 3268 | hCoV-19/USA/MN-MDH-1213/2020\|EPI_ISL_477302\|2020-0 | ···················· | ···················· | ···················· | ···················· | ···················· | ···················· | |
| D | 3269 | hCoV-19/USA/MN-MDH-1218/2020\|EPI_ISL_477307\|2020-0 | ···················· | ···················· | ···················· | ···················· | ···················· | ···················· | |
| D | 3270 | hCoV-19/USA/MN-MDH-1219/2020\|EPI_ISL_477308\|2020-0 | ···················· | ···················· | ···················· | ···················· | ···················· | ···················· | |
| D | 3271 | hCoV-19/USA/MN-MDH-1220/2020\|EPI_ISL_477309\|2020-0 | ···················· | ···················· | ···················· | ···················· | ···················· | ···················· | |
| D | 3272 | hCoV-19/England/LOND-D707B/2020\|EPI_ISL_478458\|202 | ···················· | ···················· | ···················· | ···················· | ···················· | ···················· | |
| D | 3273 | hCoV-19/England/LOND-D71E1/2020\|EPI_ISL_478461\|202 | ···················· | ···················· | ···················· | ···················· | ···················· | ···················· | |
| D | 3274 | hCoV-19/England/LOND-D71F0/2020\|EPI_ISL_478462\|202 | ···················· | ···················· | ···················· | ···················· | ···················· | ···················· | |
| D | 3275 | hCoV-19/England/LOND-D720C/2020\|EPI_ISL_478463\|202 | ···················· | ···················· | ···················· | ···················· | ···················· | ···················· | |
| D | 3276 | hCoV-19/England/LOND-D7239/2020\|EPI_ISL_478464\|202 | ···················· | ···················· | ···················· | ···················· | ···················· | ···················· | |
| D | 3277 | hCoV-19/England/LOND-D7248/2020\|EPI_ISL_478465\|202 | ···················· | ···················· | ···················· | ···················· | ···················· | ···················· | |
| D | 3278 | hCoV-19/England/LOND-D7257/2020\|EPI_ISL_478466\|202 | ···················· | ···················· | ···················· | ···················· | ···················· | ···················· | |
| D | 3279 | hCoV-19/England/LOND-D7275/2020\|EPI_ISL_478468\|202 | ···················· | ···················· | ···················· | ···················· | ···················· | ···················· | |
| D | 3280 | hCoV-19/England/LOND-D72A2/2020\|EPI_ISL_478469\|202 | ···················· | ···················· | ···················· | ···················· | ···················· | ···················· | |
| D | 3281 | hCoV-19/England/LOND-D72C0/2020\|EPI_ISL_478470\|202 | ···················· | ···················· | ···················· | ···················· | ···················· | ···················· | |
| D | 3282 | hCoV-19/England/NORT-294DCC/2020\|EPI_ISL_478614\|20 | ···················· | ···················· | ···················· | ···················· | ···················· | ···················· | |
| D | 3283 | hCoV-19/England/NORT-294DEA/2020\|EPI_ISL_478616\|20 | ···················· | ···················· | ···················· | ···················· | ···················· | ···················· | |
| D | 3284 | hCoV-19/England/NORT-294E14/2020\|EPI_ISL_478619\|20 | ···················· | ···················· | ···················· | ···················· | ···················· | ···················· | |
| D | 3285 | hCoV-19/England/NORT-294E32/2020\|EPI_ISL_478621\|20 | ···················· | ···················· | ···················· | ···················· | ···················· | ···················· | |
| D | 3286 | hCoV-19/England/NORT-294EE7/2020\|EPI_ISL_478622\|20 | ···················· | ···················· | ···················· | ···················· | ···················· | ···················· | |
| D | 3287 | hCoV-19/England/NORT-294F7B/2020\|EPI_ISL_478623\|20 | ···················· | ···················· | ···················· | ···················· | ···················· | ···················· | |
| D | 3288 | hCoV-19/England/NORT-294FF3/2020\|EPI_ISL_478624\|20 | ···················· | ···················· | ···················· | ···················· | ···················· | ···················· | |
| D | 3289 | hCoV-19/England/NORT-29501D/2020\|EPI_ISL_478626\|20 | ···················· | ···················· | ···················· | ···················· | ···················· | ···················· | |
| D | 3290 | hCoV-19/England/NORT-29502C/2020\|EPI_ISL_478627\|20 | ···················· | ···················· | ···················· | ···················· | ···················· | ···················· | |
| D | 3291 | hCoV-19/England/NORT-29503B/2020\|EPI_ISL_478628\|20 | ···················· | ···················· | ···················· | ···················· | ···················· | ···················· | |
| D | 3292 | hCoV-19/England/NORT-295068/2020\|EPI_ISL_478630\|20 | ···················· | ···················· | ···················· | ···················· | ···················· | ···················· | |
| D | 3293 | hCoV-19/England/NORT-295147/2020\|EPI_ISL_478632\|20 | ···················· | ···················· | ···················· | ···················· | ···················· | ···················· | |
| D | 3294 | hCoV-19/England/NORT-295675/2020\|EPI_ISL_478638\|20 | ···················· | ···················· | ···················· | ···················· | ···················· | ···················· | |
| D | 3295 | hCoV-19/England/NORT-295BFE/2020\|EPI_ISL_478657\|20 | ···················· | ···················· | ···················· | ···················· | ···················· | ···················· | |
| D | 3296 | hCoV-19/England/NORT-295C19/2020\|EPI_ISL_478658\|20 | ···················· | ···················· | ···················· | ···················· | ···················· | ···················· | |
| D | 3297 | hCoV-19/England/NORT-295C46/2020\|EPI_ISL_478661\|20 | ···················· | ···················· | ···················· | ···················· | ···················· | ···················· | |
| D | 3298 | hCoV-19/England/NORT-295D52/2020\|EPI_ISL_478662\|20 | ···················· | ···················· | ···················· | ···················· | ···················· | ···················· | |
| D | 3299 | hCoV-19/Turkey/KOC-IST-B303/2020\|EPI_ISL_478669\|20 | ···················· | ···················· | ···················· | ···················· | ···················· | ···················· | |
| D | 3300 | hCoV-19/Turkey/KOC-IST-B91/2020\|EPI_ISL_478670\|202 | ···················· | ···················· | ···················· | ···················· | ···················· | ···················· | |
| D | 3301 | hCoV-19/Egypt/CUNCI-HGC6I010/2020\|EPI_ISL_478672\|2 | ···················· | ···················· | ···················· | ···················· | ···················· | ···················· | |
| D | 3302 | hCoV-19/Australia/NSW561/2020\|EPI_ISL_478673\|2020- | ···················· | ···················· | ···················· | ···················· | ···················· | ···················· | |
| D | 3303 | hCoV-19/Australia/NSW562/2020\|EPI_ISL_478674\|2020- | ···················· | ···················· | ···················· | ···················· | ···················· | ···················· | |
| D | 3304 | hCoV-19/Australia/NSW563/2020\|EPI_ISL_478675\|2020- | ··················T· | ···················· | ···················· | ···················· | ···················· | ···················· | 1 |
| D | 3305 | hCoV-19/Australia/NSW564/2020\|EPI_ISL_478676\|2020- | ···················· | ···················· | ···················· | ···················· | ···················· | ···················· | |
| D | 3306 | hCoV-19/Australia/NSW565/2020\|EPI_ISL_478677\|2020- | ···················· | ···················· | ···················· | ···················· | ···················· | ···················· | |
| D | 3307 | hCoV-19/Australia/NSW566/2020\|EPI_ISL_478678\|2020- | ···················· | ···················· | ···················· | ···················· | ···················· | ···················· | |
| D | 3308 | hCoV-19/Australia/NSW567/2020\|EPI_ISL_478679\|2020- | ···················· | ···················· | ···················· | ···················· | ···················· | ···················· | |
| D | 3309 | hCoV-19/Australia/NSW568/2020\|EPI_ISL_478680\|2020- | ···················· | ···················· | ···················· | ···················· | ···················· | ···················· | |
| D | 3310 | hCoV-19/Australia/NSW569/2020\|EPI_ISL_478681\|2020- | ···················· | ···················· | ···················· | ···················· | ···················· | ···················· | |
| D | 3311 | hCoV-19/Australia/NSW570/2020\|EPI_ISL_478682\|2020- | ···················· | ···················· | ···················· | ···················· | ···················· | ···················· | |

| | | | | | | | | |
|---|---|---|---|---|---|---|---|---|
| D | 3312 | hCoV-19/Australia/NSW571/2020\|EPI_ISL_478683\|2020- | ···················· | ···················· | ···················· | ···················· | ···················· | ···················· |
| D | 3313 | hCoV-19/Australia/NSW572/2020\|EPI_ISL_478684\|2020- | ···················· | ···················· | ···················· | ···················· | ···················· | ···················· |
| D | 3314 | hCoV-19/Australia/NSW573/2020\|EPI_ISL_478685\|2020- | ···················· | ···················· | ···················· | ···················· | ···················· | ···················· |
| D | 3315 | hCoV-19/Australia/NSW574/2020\|EPI_ISL_478686\|2020- | ···················· | ···················· | ···················· | ···················· | ···················· | ···················· |
| D | 3316 | hCoV-19/Australia/NSW575/2020\|EPI_ISL_478687\|2020- | ···················· | ···················· | ···················· | ···················· | ···················· | ···················· |
| D | 3317 | hCoV-19/Australia/NSW576/2020\|EPI_ISL_478688\|2020- | ···················· | ···················· | ···················· | ···················· | ···················· | ···················· |
| D | 3318 | hCoV-19/Australia/NSW577/2020\|EPI_ISL_478689\|2020- | ···················· | ···················· | ···················· | ···················· | ···················· | ···················· |
| D | 3319 | hCoV-19/Australia/NSW578/2020\|EPI_ISL_478690\|2020- | ···················· | ···················· | ···················· | ···················· | ···················· | ···················· |
| D | 3320 | hCoV-19/Australia/NSW579/2020\|EPI_ISL_478691\|2020- | ···················· | ···················· | ···················· | ···················· | ···················· | ···················· |
| D | 3321 | hCoV-19/Australia/NSW580/2020\|EPI_ISL_478692\|2020- | ···················· | ···················· | ···················· | ···················· | ···················· | ···················· |
| D | 3322 | hCoV-19/Australia/NSW581/2020\|EPI_ISL_478693\|2020- | ···················· | ···················· | ···················· | ···················· | ···················· | ···················· |
| D | 3323 | hCoV-19/Australia/NSW582/2020\|EPI_ISL_478694\|2020- | ···················· | ···················· | ···················· | ···················· | ···················· | ···················· |
| D | 3324 | hCoV-19/Australia/NSW583/2020\|EPI_ISL_478695\|2020- | ···················· | ···················· | ···················· | ···················· | ···················· | ···················· |
| D | 3325 | hCoV-19/Australia/NSW584/2020\|EPI_ISL_478696\|2020- | ···················· | ···················· | ···················· | ···················· | ···················· | ···················· |
| D | 3326 | hCoV-19/Australia/NSW585/2020\|EPI_ISL_478697\|2020- | ···················· | ···················· | ···················· | ···················· | ···················· | ···················· |
| D | 3327 | hCoV-19/Australia/NSW586/2020\|EPI_ISL_478698\|2020- | ···················· | ···················· | ···················· | ···················· | ···················· | ···················· |
| D | 3328 | hCoV-19/Australia/NSW587/2020\|EPI_ISL_478699\|2020- | ···················· | ···················· | ···················· | ···················· | ···················· | ···················· |
| D | 3329 | hCoV-19/Canada/ON-UHTC_0033/2020\|EPI_ISL_464016\|20 | ···················· | ···················· | ···················· | ···················· | ···················· | ···················· |
| D | 3330 | hCoV-19/Canada/ON-UHTC_0038/2020\|EPI_ISL_464018\|20 | ···················· | ···················· | ···················· | ···················· | ···················· | ···················· |
| D | 3331 | hCoV-19/Canada/ON-UHTC_0076/2020\|EPI_ISL_464019\|20 | ···················· | ···················· | ···················· | ···················· | ···················· | ···················· |
| D | 3332 | hCoV-19/Canada/ON-UHTC_0043/2020\|EPI_ISL_464020\|20 | ···················· | ···················· | ···················· | ···················· | ···················· | ···················· |
| D | 3333 | hCoV-19/Canada/ON-UHTC_0069/2020\|EPI_ISL_464021\|20 | ···················· | ···················· | ···················· | ···················· | ···················· | ···················· |
| D | 3334 | hCoV-19/Canada/ON-UHTC_0070/2020\|EPI_ISL_464022\|20 | ···················· | ···················· | ···················· | ···················· | ···················· | ···················· |
| D | 3335 | hCoV-19/Canada/ON-UHTC_0058/2020\|EPI_ISL_464024\|20 | ···················· | ···················· | ···················· | ···················· | ···················· | ···················· |
| D | 3336 | hCoV-19/Canada/ON-UHTC_0039/2020\|EPI_ISL_464026\|20 | ···················· | ···················· | ···················· | ···················· | ···················· | ···················· |
| D | 3337 | hCoV-19/Canada/ON-UHTC_0037/2020\|EPI_ISL_464028\|20 | ···················· | ···················· | ···················· | ···················· | ···················· | ···················· |
| D | 3338 | hCoV-19/Canada/ON-UHTC_0085/2020\|EPI_ISL_464029\|20 | ···················· | ···················· | ···················· | ···················· | ···················· | ···················· |
| D | 3339 | hCoV-19/Canada/ON-UHTC_0065/2020\|EPI_ISL_464030\|20 | ···················· | ···················· | ···················· | ···················· | ···················· | ···················· |
| D | 3340 | hCoV-19/Canada/ON-UHTC_0035/2020\|EPI_ISL_464033\|20 | ···················· | ···················· | ···················· | ···················· | ···················· | ···················· |
| D | 3341 | hCoV-19/Canada/ON-UHTC_0050/2020\|EPI_ISL_464036\|20 | ···················· | ···················· | ···················· | ···················· | ···················· | ···················· |
| D | 3342 | hCoV-19/England/20170048204/2020\|EPI_ISL_465300\|20 | ···················· | ···················· | ···················· | ···················· | ···················· | ···················· |
| D | 3343 | hCoV-19/England/20172045704/2020\|EPI_ISL_465347\|20 | ···················· | ···················· | ···················· | ···················· | ···················· | ·············NN····· |
| D | 3344 | hCoV-19/England/20172045804/2020\|EPI_ISL_465348\|20 | ···················· | ···················· | ···················· | ···················· | ···················· | ···················· |
| D | 3345 | hCoV-19/England/20172045904/2020\|EPI_ISL_465349\|20 | ···················· | ···················· | ···················· | ···················· | ···················· | ···················· |
| D | 3346 | hCoV-19/England/20172046004/2020\|EPI_ISL_465350\|20 | ···················· | ···················· | ···················· | ···················· | ···················· | ···················· |
| D | 3347 | hCoV-19/England/20172046404/2020\|EPI_ISL_465351\|20 | ···················· | ···················· | ···················· | ···················· | ···················· | ···················· |
| D | 3348 | hCoV-19/England/20172047204/2020\|EPI_ISL_465352\|20 | ···················· | ···················· | ···················· | ···················· | ···················· | ···················· |
| D | 3349 | hCoV-19/England/20172048304/2020\|EPI_ISL_465353\|20 | ···················· | ···················· | ···················· | ···················· | ···················· | ···················· |
| D | 3350 | hCoV-19/England/20172054604/2020\|EPI_ISL_465357\|20 | ···················· | ···················· | ···················· | ···················· | ···················· | ···················· |
| D | 3351 | hCoV-19/England/20172055004/2020\|EPI_ISL_465359\|20 | ···················· | ···················· | ···················· | ···················· | ···················· | ···················· |
| D | 3353 | hCoV-19/England/20168006104/2020\|EPI_ISL_466601\|20 | ···················· | ···················· | ···················· | ···················· | ···················· | ···················· |
| D | 3354 | hCoV-19/England/20168026204/2020\|EPI_ISL_466603\|20 | ···················· | ···················· | ···················· | ···················· | ···················· | ···················· |
| D | 3357 | hCoV-19/England/20168032004/2020\|EPI_ISL_466606\|20 | ···················· | ···················· | ···················· | ···················· | ···················· | ···················· |
| D | 3358 | hCoV-19/England/20168032404/2020\|EPI_ISL_466608\|20 | ···················· | ···················· | ···················· | ···················· | ···················· | ···················· |
| D | 3360 | hCoV-19/England/20168035004/2020\|EPI_ISL_466611\|20 | ···················· | ···················· | ···················· | ···················· | ···················· | ···················· |
| D | 3361 | hCoV-19/England/20168035104/2020\|EPI_ISL_466612\|20 | ···················· | ···················· | ···················· | ···················· | ···················· | ···················· |
| D | 3362 | hCoV-19/England/20168035204/2020\|EPI_ISL_466613\|20 | ···················· | ···················· | ···················· | ···················· | ···················· | ···················· |
| D | 3363 | hCoV-19/England/20168037604/2020\|EPI_ISL_466615\|20 | ···················· | ···················· | ···················· | ···················· | ···················· | ···················· |
| D | 3364 | hCoV-19/England/20168038004/2020\|EPI_ISL_466616\|20 | ···················· | ···················· | ···················· | ···················· | ···················· | ···················· |
| D | 3365 | hCoV-19/England/20168038104/2020\|EPI_ISL_466617\|20 | ···················· | ···················· | ···················· | ···················· | ···················· | ···················· |
| D | 3367 | hCoV-19/England/20168038304/2020\|EPI_ISL_466619\|20 | ···················· | ···················· | ···················· | ···················· | ···················· | ···················· |
| D | 3369 | hCoV-19/England/20168042704/2020\|EPI_ISL_466622\|20 | ···················· | ···················· | ···················· | ···················· | ···················· | ···················· |
| D | 3370 | hCoV-19/England/20169000204/2020\|EPI_ISL_466623\|20 | ···················· | ···················· | ···················· | ···················· | ···················· | ···················· |
| D | 3372 | hCoV-19/England/20169000804/2020\|EPI_ISL_466625\|20 | ···················· | ···················· | ···················· | ···················· | ···················· | ···················· |
| D | 3373 | hCoV-19/Bangladesh/BCSIR-NILMRC-051/2020\|EPI_ISL_4 | ···················· | ···················· | ···················· | ···················· | ···················· | ···················· |
| D | 3374 | hCoV-19/Bangladesh/BCSIR-NILMRC-052/2020\|EPI_ISL_4 | ···················· | ···················· | ···················· | ···················· | ···················· | ···················· |
| D | 3375 | hCoV-19/Bangladesh/BCSIR-NILMRC-054/2020\|EPI_ISL_4 | ···················· | ···················· | ···················· | ···················· | ···················· | ···················· |
| D | 3376 | hCoV-19/Bangladesh/BCSIR-NILMRC-056/2020\|EPI_ISL_4 | ···················· | ···················· | ···················· | ···················· | ···················· | ···················· |
| D | 3377 | hCoV-19/Bangladesh/BCSIR-NILMRC-057/2020\|EPI_ISL_4 | ···················· | ···················· | ···················· | ···················· | ···················· | ···················· |
| D | 3379 | hCoV-19/USA/NE-11510/2020\|EPI_ISL_466678\|2020-05-0 | ···················· | ················T··· | ···················· | ···················· | ···················· | ···················· |
| D | 3380 | hCoV-19/USA/NE-11647/2020\|EPI_ISL_466680\|2020-05-0 | ···················· | ···················· | ···················· | ···················· | ···················· | ···················· |
| D | 3381 | hCoV-19/Bangladesh/BCSIR-NILMRC-088/2020\|EPI_ISL_4 | ···················· | ···················· | ···················· | ···················· | ···················· | ···················· |
| D | 3382 | hCoV-19/Bangladesh/BCSIR-NILMRC-072/2020\|EPI_ISL_4 | ···················· | ···················· | ···················· | ···················· | ···················· | ···················· |
| D | 3383 | hCoV-19/Bangladesh/BCSIR-NILMRC-086/2020\|EPI_ISL_4 | ···················· | ···················· | ···················· | ···················· | ···················· | ···················· |
| D | 3384 | hCoV-19/Bangladesh/BCSIR-NILMRC-073/2020\|EPI_ISL_4 | ···················· | ···················· | ···················· | ···················· | ···················· | ···················· |
| D | 3385 | hCoV-19/Bangladesh/BCSIR-NILMRC-074/2020\|EPI_ISL_4 | ···················· | ···················· | ···················· | ···················· | ···················· | ···················· |
| D | 3386 | hCoV-19/Canada/BC_00576317/2020\|EPI_ISL_466696\|202 | ···················· | ···················· | ···················· | ···················· | ···················· | ···················· |
| D | 3387 | hCoV-19/Canada/BC_00646519/2020\|EPI_ISL_466697\|202 | ···················· | ···················· | ···················· | ···················· | ···················· | ···················· |
| D | 3388 | hCoV-19/Canada/BC_00655815/2020\|EPI_ISL_466698\|202 | ···················· | ···················· | ···················· | ···················· | ···················· | ···················· |
| D | 3389 | hCoV-19/USA/NY-QDX-42/2020\|EPI_ISL_467900\|2020-03- | ···················· | ···················· | ···················· | ···················· | ···················· | ···················· |
| D | 3390 | hCoV-19/USA/NY-QDX-72/2020\|EPI_ISL_467901\|2020-03- | ···················· | ···················· | ···················· | ···················· | ···················· | ···················· |
| D | 3391 | hCoV-19/USA/NY-QDX-75/2020\|EPI_ISL_467902\|2020-03- | ···················· | ···················· | ···················· | ···················· | ···················· | ···················· |
| D | 3392 | hCoV-19/USA/NY-QDX-81/2020\|EPI_ISL_467903\|2020-03- | ···················· | ···················· | ···················· | ···················· | ···················· | ···················· |
| D | 3393 | hCoV-19/USA/PA-QDX-30/2020\|EPI_ISL_467904\|2020-03- | ···················· | ···················· | ···················· | ···················· | ···················· | ···················· |
| D | 3394 | hCoV-19/USA/TN-QDX-126/2020\|EPI_ISL_467905\|2020-03 | ···················· | ···················· | ···················· | ···················· | ···················· | ···················· |
| D | 3395 | hCoV-19/USA/TX-QDX-25/2020\|EPI_ISL_467906\|2020-03- | ···················· | ···················· | ···················· | ···················· | ···················· | ···················· |
| D | 3396 | hCoV-19/USA/TX-QDX-37/2020\|EPI_ISL_467907\|2020-03- | ···················· | ···················· | ···················· | ···················· | ···················· | ···················· |
| D | 3397 | hCoV-19/USA/TX-QDX-60/2020\|EPI_ISL_467908\|2020-03- | ···················· | ···················· | ···················· | ···················· | ···················· | ···················· |

| | | | | | | | |
|---|---|---|---|---|---|---|---|
| D | 3398 hCoV-19/USA/UT-QDX-118/2020\|EPI_ISL_467909\|2020-03 | ···················· | ···················· | ···················· | ···················· | ···················· | ···················· |
| D | 3399 hCoV-19/USA/UT-QDX-119/2020\|EPI_ISL_467910\|2020-03 | ···················· | ···················· | ···················· | ···················· | ···················· | ···················· |
| D | 3400 hCoV-19/USA/UT-QDX-120/2020\|EPI_ISL_467911\|2020-03 | ···················· | ···················· | ···················· | ···················· | ···················· | ···················· |
| D | 3401 hCoV-19/USA/UT-QDX-131/2020\|EPI_ISL_467912\|2020-03 | ···················· | ···················· | ···················· | ···················· | ···················· | ···················· |
| D | 3402 hCoV-19/USA/UT-QDX-132/2020\|EPI_ISL_467913\|2020-03 | ···················· | ···················· | ···················· | ···················· | ···················· | ···················· |
| D | 3403 hCoV-19/USA/UT-QDX-151/2020\|EPI_ISL_467914\|2020-03 | ···················· | ···················· | ···················· | ···················· | ···················· | ···················· |
| D | 3404 hCoV-19/USA/UT-QDX-17/2020\|EPI_ISL_467915\|2020-03- | ···················· | ···················· | ···················· | ···················· | ···················· | ···················· |
| D | 3405 hCoV-19/USA/UT-QDX-18/2020\|EPI_ISL_467916\|2020-03- | ···················· | ···················· | ···················· | ···················· | ···················· | ···················· |
| D | 3406 hCoV-19/USA/UT-QDX-20/2020\|EPI_ISL_467917\|2020-03- | ···················· | ···················· | ···················· | ···················· | ···················· | ···················· |
| D | 3407 hCoV-19/USA/UT-QDX-27/2020\|EPI_ISL_467918\|2020-03- | ···················· | ···················· | ···················· | ···················· | ···················· | ···················· |
| D | 3408 hCoV-19/USA/UT-QDX-49/2020\|EPI_ISL_467919\|2020-03- | ···················· | ···················· | ···················· | ···················· | ···················· | ···················· |
| D | 3409 hCoV-19/USA/UT-QDX-57/2020\|EPI_ISL_467920\|2020-03- | ···················· | ···················· | ···················· | ···················· | ···················· | ···················· |
| D | 3410 hCoV-19/USA/UT-QDX-63/2020\|EPI_ISL_467921\|2020-03- | ···················· | ···················· | ···················· | ···················· | ···················· | ···················· |
| D | 3411 hCoV-19/USA/UT-QDX-65/2020\|EPI_ISL_467922\|2020-03- | ···················· | ···················· | ···················· | ···················· | ···················· | ···················· |
| D | 3412 hCoV-19/USA/UT-QDX-66/2020\|EPI_ISL_467923\|2020-03- | ···················· | ···················· | ···················· | ···················· | ···················· | ···················· |
| D | 3413 hCoV-19/USA/UT-QDX-77/2020\|EPI_ISL_467924\|2020-03- | ···················· | ···················· | ···················· | ···················· | ···················· | ···················· |
| D | 3414 hCoV-19/USA/UT-QDX-82/2020\|EPI_ISL_467925\|2020-03- | ···················· | ···················· | ···················· | ···················· | ···················· | ···················· |
| D | 3415 hCoV-19/USA/WA-QDX-22/2020\|EPI_ISL_467926\|2020-03- | ···················· | ···················· | ···················· | ···················· | ···················· | ···················· |
| D | 3416 hCoV-19/USA/WA-QDX-62/2020\|EPI_ISL_467927\|2020-03- | ···················· | ···················· | ···················· | ···················· | ···················· | ···················· |
| D | 3417 hCoV-19/USA/VA-DCLS-0315/2020\|EPI_ISL_467928\|2020- | ···················· | ···················· | ···················· | ···················· | ···················· | ···················· |
| D | 3418 hCoV-19/USA/VA-DCLS-0316/2020\|EPI_ISL_467929\|2020- | ···················· | ···················· | ···················· | ···················· | ···················· | ···················· |
| D | 3419 hCoV-19/USA/VA-DCLS-0317/2020\|EPI_ISL_467930\|2020- | ···················· | ···················· | ···················· | ···················· | ···················· | ···················· |
| D | 3420 hCoV-19/USA/VA-DCLS-0321/2020\|EPI_ISL_467932\|2020- | ···················· | ···················· | ···················· | ···················· | ···················· | ···················· |
| D | 3421 hCoV-19/USA/VA-DCLS-0322/2020\|EPI_ISL_467933\|2020- | ···················· | ···················· | ···················· | ···················· | ···················· | ···················· |
| D | 3422 hCoV-19/USA/VA-DCLS-0329/2020\|EPI_ISL_467934\|2020- | ···················· | ···················· | ···················· | ···················· | ···················· | ···················· |
| D | 3423 hCoV-19/USA/VA-DCLS-0334/2020\|EPI_ISL_467936\|2020- | ···················· | ···················· | ···················· | ···················· | ···················· | ···················· |
| D | 3424 hCoV-19/USA/VA-DCLS-0335/2020\|EPI_ISL_467937\|2020- | ···················· | ···················· | ···················· | ···················· | ···················· | ···················· |
| D | 3425 hCoV-19/USA/VA-DCLS-0336/2020\|EPI_ISL_467938\|2020- | ···················· | ···················· | ···················· | ···················· | ···················· | ···················· |
| D | 3426 hCoV-19/USA/VA-DCLS-0367/2020\|EPI_ISL_467941\|2020- | ···················· | ···················· | ···················· | ···················· | ···················· | ···················· |
| D | 3427 hCoV-19/USA/VA-DCLS-0368/2020\|EPI_ISL_467942\|2020- | ···················· | ···················· | ···················· | ···················· | ···················· | ···················· |
| D | 3428 hCoV-19/USA/VA-DCLS-0382/2020\|EPI_ISL_467943\|2020- | ···················· | ···················· | ···················· | ···················· | ···················· | ···················· |
| D | 3429 hCoV-19/USA/VA-DCLS-0384/2020\|EPI_ISL_467944\|2020- | ···················· | ···················· | ···················· | ···················· | ···················· | ···················· |
| D | 3430 hCoV-19/USA/NY-AECOM-036/2020\|EPI_ISL_467945\|2020- | ···················· | ···················· | ···················· | ···················· | ···················· | ···················· |
| D | 3431 hCoV-19/USA/CA-IGI-0006/2020\|EPI_ISL_467946\|2020-0 | ···················· | ···················· | ···················· | ···················· | ···················· | ···················· |
| D | 3432 hCoV-19/USA/CA-IGI-0031/2020\|EPI_ISL_467948\|2020-0 | ···················· | ···················· | ···················· | ···················· | ···················· | ···················· |
| D | 3433 hCoV-19/USA/CA-SR0415/2020\|EPI_ISL_467950\|2020-03- | ···················· | ···················· | ···················· | ···················· | ···················· | ···················· |
| D | 3434 hCoV-19/USA/CA-SR0416/2020\|EPI_ISL_467951\|2020-03- | ···················· | ···················· | ···················· | ···················· | ···················· | ···················· |
| D | 3435 hCoV-19/USA/CA-SR0418/2020\|EPI_ISL_467952\|2020-03- | ···················· | ···················· | ···················· | ···················· | ···················· | ···················· |
| D | 3436 hCoV-19/USA/CA-SR0419/2020\|EPI_ISL_467953\|2020-03- | ···················· | ·A·················· | ···················· | ···················· | ···················· | ···················· |
| D | 3437 hCoV-19/USA/CA-SR0421/2020\|EPI_ISL_467954\|2020-03- | ···················· | ···················· | ···················· | ···················· | ···················· | ···················· |
| D | 3438 hCoV-19/USA/CA-SR0424/2020\|EPI_ISL_467955\|2020-03- | ···················· | ···················· | ···················· | ···················· | ···················· | ···················· |
| D | 3439 hCoV-19/USA/CA-SR0425/2020\|EPI_ISL_467956\|2020-03- | ···················· | ···················· | ···················· | ···················· | ···················· | ···················· |
| D | 3440 hCoV-19/USA/CA-SR0426/2020\|EPI_ISL_467957\|2020-03- | ···················· | ···················· | ···················· | ···················· | ···················· | ···················· |
| D | 3441 hCoV-19/USA/CA-SR0428/2020\|EPI_ISL_467958\|2020-03- | ···················· | ···················· | ···················· | ···················· | ···················· | ···················· |
| D | 3442 hCoV-19/USA/CA-SR0429/2020\|EPI_ISL_467959\|2020-03- | ···················· | ···················· | ···················· | ···················· | ···················· | ···················· |
| D | 3443 hCoV-19/USA/CA-SR0430/2020\|EPI_ISL_467960\|2020-03- | ···················· | ···················· | ···················· | ···················· | ···················· | ···················· |
| D | 3444 hCoV-19/USA/CA-SR0431/2020\|EPI_ISL_467961\|2020-03- | ···················· | ···················· | ···················· | ···················· | ···················· | ···················· |
| D | 3445 hCoV-19/USA/CA-SR0432/2020\|EPI_ISL_467962\|2020-03- | ···················· | ···················· | ···················· | ···················· | ···················· | ···················· |
| D | 3446 hCoV-19/USA/CA-SR0434/2020\|EPI_ISL_467963\|2020-04- | ···················· | ···················· | ···················· | ···················· | ···················· | ···················· |
| D | 3447 hCoV-19/USA/MN-GMF-05101/2020\|EPI_ISL_471144\|2020- | ···················· | ···················· | ···················· | ···················· | ···················· | ···················· |
| D | 3448 hCoV-19/USA/WI-GMF-05143/2020\|EPI_ISL_471145\|2020- | ···················· | ···················· | ···················· | ···················· | ···················· | ···················· |
| D | 3449 hCoV-19/USA/WI-GMF-05510/2020\|EPI_ISL_471146\|2020- | ···················· | ·············A······ | ···················· | ···················· | ···················· | ···················· |
| D | 3450 hCoV-19/USA/WI-GMF-05641/2020\|EPI_ISL_471147\|2020- | ···················· | ···················· | ···················· | ···················· | ···················· | ···················· |
| D | 3451 hCoV-19/USA/WI-GMF-05642/2020\|EPI_ISL_471148\|2020- | ···················· | ···················· | ···················· | ···················· | ···················· | ···················· |
| D | 3452 hCoV-19/USA/WI-GMF-05678/2020\|EPI_ISL_471149\|2020- | ···················· | ···················· | ···················· | ···················· | ···················· | ···················· |
| D | 3453 hCoV-19/USA/WI-GMF-05945/2020\|EPI_ISL_471150\|2020- | ···················· | ···················· | ···················· | ···················· | ···················· | ···················· |
| D | 3454 hCoV-19/USA/WI-GMF-06001/2020\|EPI_ISL_471151\|2020- | ···················· | ···················· | ···················· | ···················· | ···················· | ···················· |
| D | 3455 hCoV-19/USA/WI-GMF-06054/2020\|EPI_ISL_471152\|2020- | ···················· | ···················· | ···················· | ···················· | ···················· | ···················· |
| D | 3456 hCoV-19/USA/WI-GMF-06127/2020\|EPI_ISL_471153\|2020- | ···················· | ···················· | ···················· | ···················· | ···················· | ···················· |
| D | 3457 hCoV-19/USA/WI-GMF-06190/2020\|EPI_ISL_471154\|2020- | ···················· | ···················· | ···················· | ···················· | ···················· | ···················· |
| D | 3458 hCoV-19/USA/WI-GMF-06192/2020\|EPI_ISL_471155\|2020- | ···················· | ···················· | ···················· | ···················· | ···················· | ···················· |
| D | 3459 hCoV-19/USA/WI-GMF-06288/2020\|EPI_ISL_471156\|2020- | ···················· | ···················· | ···················· | ···················· | ···················· | ···················· |
| D | 3460 hCoV-19/USA/WI-GMF-M00007/2020\|EPI_ISL_471157\|2020 | ···················· | ···················· | ···················· | ···················· | ···················· | ···················· |
| D | 3461 hCoV-19/Gambia/0214/2020\|EPI_ISL_471158\|2020-03-29 | ···················· | ···················· | ···················· | ···················· | ···················· | ···················· |
| D | 3462 hCoV-19/Gambia/0536/2020\|EPI_ISL_471163\|2020-04-30 | ···················· | ···················· | ···················· | ···················· | ···················· | ···················· |
| D | 3463 hCoV-19/Gambia/0547/2020\|EPI_ISL_471164\|2020-04-30 | ···················· | ···················· | ···················· | ···················· | ···················· | ···················· |
| D | 3464 hCoV-19/Gambia/1094/2020\|EPI_ISL_471167\|2020-05-04 | ···················· | ···················· | ···················· | ···················· | ···················· | ···················· |
| D | 3465 hCoV-19/Gambia/1277/2020\|EPI_ISL_471171\|2020-05-07 | ···················· | ···················· | ···················· | ···················· | ···················· | ···················· |
| D | 3466 hCoV-19/Norway/2454/2020\|EPI_ISL_471172\|2020-04-17 | ···················· | ···················· | ···················· | ···················· | ···················· | ···················· |
| D | 3467 hCoV-19/Norway/2466/2020\|EPI_ISL_471173\|2020-04-21 | ···················· | ···················· | ···················· | ···················· | ···················· | ···················· |
| D | 3468 hCoV-19/Norway/2534/2020\|EPI_ISL_471176\|2020-05-04 | ···················· | ···················· | ···················· | ···················· | ···················· | ···················· |
| D | 3469 hCoV-19/Norway/2549/2020\|EPI_ISL_471177\|2020-05-04 | ···················· | ···················· | ···················· | ···················· | ···················· | ···················· |
| D | 3470 hCoV-19/USA/WI-WSLH-200012/2020\|EPI_ISL_471178\|202 | ···················· | ···················· | ···················· | ···················· | ···················· | ···················· |
| D | 3471 hCoV-19/USA/WI-WSLH-200014/2020\|EPI_ISL_471180\|202 | ···················· | ···················· | ···················· | ···················· | ···················· | ···················· |
| D | 3472 hCoV-19/USA/WI-WSLH-200015/2020\|EPI_ISL_471181\|202 | ···················· | ···················· | ···················· | ···················· | ···················· | ···················· |
| D | 3473 hCoV-19/USA/WI-WSLH-200017/2020\|EPI_ISL_471183\|202 | ···················· | ···················· | ···················· | ···················· | ···················· | ···················· |
| D | 3474 hCoV-19/USA/WI-WSLH-200019/2020\|EPI_ISL_471185\|202 | ···················· | ···················· | ···················· | ···················· | ···················· | ···················· |
| D | 3475 hCoV-19/USA/WI-WSLH-200021/2020\|EPI_ISL_471187\|202 | ···················· | ···················· | ···················· | ···················· | ···················· | ···················· |

| | | | | | | | | |
|---|---|---|---|---|---|---|---|---|
| D | 3476 hCoV-19/USA/WI-WSLH-200023/2020\|EPI_ISL_471188\|202 | ···················· | ···················· | ···················· | ···················· | ···················· | ···················· | |
| D | 3477 hCoV-19/USA/WI-WSLH-200024/2020\|EPI_ISL_471189\|202 | ···················· | ···················· | ···················· | ···················· | ···················· | ···················· | |
| D | 3478 hCoV-19/USA/WI-WSLH-200025/2020\|EPI_ISL_471190\|202 | ···················· | ···················· | ···················· | ···················· | ···················· | ···················· | |
| D | 3479 hCoV-19/USA/WI-WSLH-200026/2020\|EPI_ISL_471191\|202 | ···················· | ···················· | ···················· | ···················· | ···················· | ···················· | |
| D | 3480 hCoV-19/USA/WI-WSLH-200027/2020\|EPI_ISL_471192\|202 | ···················· | ···················· | ···················· | ···················· | ···················· | ···················· | |
| D | 3481 hCoV-19/USA/WI-WSLH-200029/2020\|EPI_ISL_471194\|202 | ···················· | ···················· | ···················· | ···················· | ···················· | ···················· | |
| D | 3482 hCoV-19/USA/WI-WSLH-200031/2020\|EPI_ISL_471196\|202 | ···················· | ···················· | ···················· | ···················· | ···················· | ···················· | |
| D | 3483 hCoV-19/USA/WI-WSLH-200032/2020\|EPI_ISL_471197\|202 | ···················· | ···················· | ···················· | ···················· | ···················· | ···················· | |
| D | 3484 hCoV-19/USA/WI-WSLH-200034/2020\|EPI_ISL_471199\|202 | ···················· | ···················· | ···················· | ···················· | ········C··········· | ···················· | |
| D | 3485 hCoV-19/England/NOTT-111D22/2020\|EPI_ISL_472400\|20 | ···················· | ···················· | ···················· | ···················· | ···················· | ···················· | |
| D | 3486 hCoV-19/England/NOTT-111D31/2020\|EPI_ISL_472401\|20 | ···················· | ···················· | ···················· | ···················· | ···················· | ···················· | |
| D | 3487 hCoV-19/England/NOTT-111D40/2020\|EPI_ISL_472402\|20 | ···················· | ···················· | ···················· | ···················· | ···················· | ···················· | |
| D | 3488 hCoV-19/England/NOTT-111D5F/2020\|EPI_ISL_472403\|20 | ···················· | ···················· | ···················· | ···················· | ···················· | ···················· | |
| D | 3489 hCoV-19/England/NOTT-111D6E/2020\|EPI_ISL_472404\|20 | ···················· | ···················· | ···················· | ···················· | ···················· | ···················· | |
| D | 3490 hCoV-19/England/NOTT-111DB9/2020\|EPI_ISL_472405\|20 | ···················· | ···················· | ···················· | ···················· | ···················· | ···················· | |
| D | 3491 hCoV-19/England/NOTT-111DC8/2020\|EPI_ISL_472406\|20 | ···················· | ···················· | ···················· | ···················· | ···················· | ···················· | |
| D | 3492 hCoV-19/England/NOTT-111DD7/2020\|EPI_ISL_472407\|20 | ···················· | ···················· | ···················· | ···················· | ···················· | ···················· | |
| D | 3493 hCoV-19/England/NOTT-111DE6/2020\|EPI_ISL_472408\|20 | ···················· | ···················· | ···················· | ···················· | ···················· | ···················· | |
| D | 3494 hCoV-19/England/NOTT-111DF5/2020\|EPI_ISL_472409\|20 | ···················· | ···················· | ···················· | ···················· | ···················· | ···················· | |
| D | 3495 hCoV-19/England/NOTT-111E01/2020\|EPI_ISL_472410\|20 | ···················· | ···················· | ···················· | ···················· | ···················· | ···················· | |
| D | 3496 hCoV-19/England/NOTT-111E10/2020\|EPI_ISL_472411\|20 | ···················· | ···················· | ···················· | ···················· | ···················· | ···················· | |
| D | 3497 hCoV-19/England/NOTT-111E2F/2020\|EPI_ISL_472412\|20 | ···················· | ···················· | ···················· | ···················· | ···················· | ···················· | |
| D | 3498 hCoV-19/England/NOTT-111E3E/2020\|EPI_ISL_472413\|20 | ···················· | ···················· | ···················· | ···················· | ···················· | ···················· | |
| D | 3499 hCoV-19/England/NOTT-111E4D/2020\|EPI_ISL_472414\|20 | ···················· | ···················· | ···················· | ···················· | ···················· | ···················· | |
| D | 3500 hCoV-19/England/NOTT-111E5C/2020\|EPI_ISL_472415\|20 | ···················· | ···················· | ···················· | ···················· | ···················· | ···················· | |
| D | 3501 hCoV-19/England/NOTT-111E7A/2020\|EPI_ISL_472416\|20 | ···················· | ···················· | ···················· | ···················· | ···················· | ···················· | |
| D | 3502 hCoV-19/England/NOTT-111E98/2020\|EPI_ISL_472418\|20 | ···················· | ···················· | ···················· | ···················· | ···················· | ···················· | |
| D | 3503 hCoV-19/England/NOTT-111EB6/2020\|EPI_ISL_472419\|20 | ···················· | ···················· | ···················· | ···················· | ···················· | ···················· | |
| D | 3504 hCoV-19/England/NOTT-111EC5/2020\|EPI_ISL_472420\|20 | ···················· | ···················· | ···················· | ···················· | ···················· | ···················· | |
| D | 3505 hCoV-19/England/NOTT-111EF2/2020\|EPI_ISL_472423\|20 | ···················· | ···················· | ···················· | ···················· | ···················· | ···················· | |
| D | 3506 hCoV-19/England/NOTT-111F1D/2020\|EPI_ISL_472424\|20 | ···················· | ···················· | ···················· | ···················· | ···················· | ···················· | |
| D | 3507 hCoV-19/England/NOTT-111F2C/2020\|EPI_ISL_472425\|20 | ···················· | ···················· | ···················· | ···················· | ···················· | ···················· | |
| D | 3508 hCoV-19/England/NOTT-111F3B/2020\|EPI_ISL_472426\|20 | ···················· | ···················· | ···················· | ···················· | ···················· | ···················· | |
| D | 3509 hCoV-19/Wales/PHWC-15C0E6/2020\|EPI_ISL_472437\|2020 | ···················· | ···················· | ···················· | ···················· | ···················· | ···················· | |
| D | 3510 hCoV-19/Wales/PHWC-15C101/2020\|EPI_ISL_472439\|2020 | ···················· | ···················· | ···················· | ···················· | ···················· | ···················· | |
| D | 3511 hCoV-19/Wales/PHWC-15C17A/2020\|EPI_ISL_472445\|2020 | ···················· | ···················· | ···················· | ···················· | ···················· | ···················· | |
| D | 3512 hCoV-19/Wales/PHWC-15C1C5/2020\|EPI_ISL_472450\|2020 | ···················· | ···················· | ···················· | ···················· | ···················· | ···················· | |
| D | 3513 hCoV-19/Wales/PHWC-15C1F2/2020\|EPI_ISL_472452\|2020 | ···················· | ···················· | ···················· | ···················· | ···················· | ···················· | |
| D | 3514 hCoV-19/Wales/PHWC-15C21D/2020\|EPI_ISL_472454\|2020 | ···················· | ···················· | ···················· | ···················· | ···················· | ···················· | |
| D | 3515 hCoV-19/Wales/PHWC-15C268/2020\|EPI_ISL_472459\|2020 | ···················· | ···················· | ···················· | ···················· | ···················· | ···················· | |
| D | 3516 hCoV-19/Wales/PHWC-15C286/2020\|EPI_ISL_472461\|2020 | ···················· | ···················· | ···················· | ···················· | ···················· | ···················· | |
| D | 3517 hCoV-19/Wales/PHWC-15C3A1/2020\|EPI_ISL_472479\|2020 | ···················· | ···················· | ···················· | ···················· | ···················· | ···················· | |
| D | 3518 hCoV-19/Wales/PHWC-15C3B0/2020\|EPI_ISL_472480\|2020 | ···················· | ···················· | ···················· | ···················· | ···················· | ···················· | |
| D | 3519 hCoV-19/Wales/PHWC-15C3DE/2020\|EPI_ISL_472482\|2020 | ···················· | ···················· | ···················· | ···················· | ···················· | ···················· | |
| D | 3520 hCoV-19/Wales/PHWC-15C453/2020\|EPI_ISL_472489\|2020 | ···················· | ···················· | ···················· | ···················· | ···················· | ···················· | |
| D | 3521 hCoV-19/Wales/PHWC-15C4F9/2020\|EPI_ISL_472498\|2020 | ···················· | ···················· | ···················· | ···················· | ···················· | ···················· | |
| D | 3522 hCoV-19/Scotland/CVR3592/2020\|EPI_ISL_473721\|2020- | ···················· | ··············C····· | ···················· | ···················· | ···················· | ···················· | |
| D | 3523 hCoV-19/Scotland/CVR3593/2020\|EPI_ISL_473722\|2020- | ···················· | ···················· | ···················· | ···················· | ···················· | ···················· | |
| D | 3524 hCoV-19/Scotland/CVR3594/2020\|EPI_ISL_473723\|2020- | ···················· | ···················· | ···················· | ···················· | ···················· | ···················· | |
| D | 3525 hCoV-19/Scotland/CVR3596/2020\|EPI_ISL_473724\|2020- | ···················· | ···················· | ···················· | ···················· | ···················· | ···················· | |
| D | 3526 hCoV-19/Scotland/CVR3598/2020\|EPI_ISL_473725\|2020- | ···················· | ·A·················· | ···················· | ···················· | ···················· | ···················· | |
| D | 3527 hCoV-19/Scotland/CVR3611/2020\|EPI_ISL_473731\|2020- | ···················· | ···················· | ···················· | ···················· | ···················· | ···················· | |
| D | 3529 hCoV-19/Scotland/CVR3614/2020\|EPI_ISL_473734\|2020- | ···················· | ···················A | ···················· | ···················· | ···················· | ···················· | 1 |
| D | 3530 hCoV-19/Scotland/CVR3630/2020\|EPI_ISL_473745\|2020- | ···················· | ···················A | ···················· | ···················· | ···················· | ···················· | 1 |
| D | 3531 hCoV-19/Scotland/CVR3632/2020\|EPI_ISL_473746\|2020- | ···················· | ···················· | ···················· | ···················· | ···················· | ···················· | |
| D | 3532 hCoV-19/Scotland/CVR3637/2020\|EPI_ISL_473748\|2020- | ···················· | ···················· | ···················· | ···················· | ···················· | ···················· | |
| D | 3533 hCoV-19/Scotland/CVR3650/2020\|EPI_ISL_473749\|2020- | ···················· | ···················· | ···················· | ···················· | ···················· | ···················· | |
| D | 3534 hCoV-19/Scotland/CVR3664/2020\|EPI_ISL_473750\|2020- | ···················· | ···················· | ···················· | ···················· | ···················· | ···················· | |
| D | 3536 hCoV-19/Scotland/CVR3698/2020\|EPI_ISL_473761\|2020- | ···················· | ·A·················· | ···················· | ···················· | ···················· | ···················· | |
| D | 3537 hCoV-19/Scotland/CVR3703/2020\|EPI_ISL_473762\|2020- | ···················· | ···················· | ···················· | ···················· | ···················· | ···················· | |
| D | 3538 hCoV-19/Scotland/CVR3709/2020\|EPI_ISL_473764\|2020- | ···················· | ···················· | ···················· | ···················· | ···················· | ···················· | |
| D | 3540 hCoV-19/Scotland/CVR3728/2020\|EPI_ISL_473768\|2020- | ···················· | ···················· | ···················· | ···················· | ···················· | ···················· | |
| D | 3541 hCoV-19/Scotland/CVR3729/2020\|EPI_ISL_473769\|2020- | ···················· | ···················· | ···················· | ···················· | ···················· | ···················· | |
| D | 3542 hCoV-19/Scotland/CVR3732/2020\|EPI_ISL_473770\|2020- | ···················· | ···················· | ···················· | ···················· | ···················· | ···················· | |
| D | 3543 hCoV-19/Scotland/CVR3733/2020\|EPI_ISL_473771\|2020- | ···················· | ···················A | ···················· | ···················· | ···················· | ···················· | 1 |
| D | 3544 hCoV-19/Scotland/CVR3735/2020\|EPI_ISL_473772\|2020- | ···················· | ···················· | ···················· | ···················· | ···················· | ···················· | |
| D | 3545 hCoV-19/Scotland/CVR3736/2020\|EPI_ISL_473773\|2020- | ···················· | ···················A | ···················· | ···················· | ···················· | ···················· | 1 |
| D | 3546 hCoV-19/Scotland/CVR3738/2020\|EPI_ISL_473774\|2020- | ···················· | ···················· | ···················· | ···················· | ···················· | ···················· | |
| D | 3547 hCoV-19/Scotland/CVR3742/2020\|EPI_ISL_473776\|2020- | ···················· | ···················A | ···················· | ···················· | ···················· | ···················· | 1 |
| D | 3548 hCoV-19/Scotland/CVR3744/2020\|EPI_ISL_473777\|2020- | ···················· | ···················A | ···················· | ···················· | ···················· | ···················· | 1 |
| D | 3549 hCoV-19/Scotland/CVR3838/2020\|EPI_ISL_473778\|2020- | ···················· | ···················· | ···················· | ···················· | ···················· | ···················· | |
| D | 3550 hCoV-19/Scotland/CVR3839/2020\|EPI_ISL_473779\|2020- | ···················· | ···················A | ···················· | ···················· | ···················· | ···················· | 1 |
| D | 3551 hCoV-19/Scotland/CVR3841/2020\|EPI_ISL_473780\|2020- | ···················· | ···················· | ···················· | ···················· | ···················· | ···················· | |
| D | 3552 hCoV-19/Scotland/CVR3842/2020\|EPI_ISL_473781\|2020- | ···················· | ···················· | ···················· | ···················· | ···················· | ···················· | |
| D | 3553 hCoV-19/Scotland/CVR991/2020\|EPI_ISL_473782\|2020-0 | ···················· | ···················· | ···················· | ···················· | ···················· | ···················· | |
| D | 3554 hCoV-19/Scotland/EDB1063/2020\|EPI_ISL_473783\|2020- | ···················· | ···················· | ···················· | ···················· | ···················· | ···················· | |
| D | 3555 hCoV-19/Scotland/EDB2245/2020\|EPI_ISL_473784\|2020- | ···················· | ···················· | ···················· | ···················· | ···················· | ···················· | |
| D | 3556 hCoV-19/Scotland/EDB2626/2020\|EPI_ISL_473785\|2020- | ···················· | ···················· | ···················· | ···················· | ···················· | ···················· | |

| | | | | | | | | | |
|---|---|---|---|---|---|---|---|---|---|
| D | 3557 | hCoV-19/Scotland/EDB2629/2020\|EPI_ISL_473786\|2020- | ···················· | ···················· | ····················· | ···················· | ···················· | ···················· | |
| D | 3558 | hCoV-19/Scotland/EDB2633/2020\|EPI_ISL_473787\|2020- | ···················· | ···················· | ····················· | ···················· | ···················· | ···················· | |
| D | 3559 | hCoV-19/Scotland/EDB2634/2020\|EPI_ISL_473788\|2020- | ···················· | ···················· | ····················· | ···················· | ···················· | ···················· | |
| D | 3560 | hCoV-19/Scotland/EDB3506/2020\|EPI_ISL_473789\|2020- | ···················· | ···················A | ····················· | ···················· | ···················· | ···················· | 1 |
| D | 3561 | hCoV-19/Scotland/EDB3654/2020\|EPI_ISL_473790\|2020- | ···················· | ···················· | ····················· | ···················· | ···················· | ···················· | |
| D | 3562 | hCoV-19/Scotland/EDB4039/2020\|EPI_ISL_473791\|2020- | ···················· | ···················· | ····················· | ···················· | ···················· | ···················· | |
| D | 3563 | hCoV-19/Scotland/EDB4041/2020\|EPI_ISL_473792\|2020- | ···················· | ···················· | ····················· | ···················· | ···················· | ···················· | |
| D | 3564 | hCoV-19/Scotland/EDB4043/2020\|EPI_ISL_473793\|2020- | ···················· | ·A·················· | ····················· | ···················· | ···················· | ···················· | |
| D | 3565 | hCoV-19/Scotland/EDB4044/2020\|EPI_ISL_473794\|2020- | ···················· | ···················· | ····················· | ···················· | ···················· | ···················· | |
| D | 3566 | hCoV-19/Scotland/EDB4471/2020\|EPI_ISL_473795\|2020- | ···················· | ···················· | ····················· | ···················· | ···················· | ···················· | |
| D | 3567 | hCoV-19/Scotland/EDB4579/2020\|EPI_ISL_473796\|2020- | ···················· | ···················· | ····················· | ···················· | ···················· | ···················· | |
| D | 3568 | hCoV-19/Scotland/EDB4584/2020\|EPI_ISL_473797\|2020- | ···················· | ···················· | ····················· | ···················· | ···················· | ···················· | |
| D | 3569 | hCoV-19/Scotland/EDB4888/2020\|EPI_ISL_473798\|2020- | ···················· | ···················· | ····················· | ···················· | ···················· | ···················· | |
| D | 3570 | hCoV-19/Scotland/EDB4928/2020\|EPI_ISL_473799\|2020- | ···················· | ···················· | ····················· | ···················· | ···················· | ···················· | |
| D | 3571 | hCoV-19/USA/MN-MDH-1214/2020\|EPI_ISL_477303\|2020-0 | ···················· | ···················· | ····················· | ···················· | ···················· | ···················· | |
| D | 3572 | hCoV-19/USA/MN-MDH-1215/2020\|EPI_ISL_477304\|2020-0 | ···················· | ···················· | ····················· | ···················· | ···················· | ···················· | |
| D | 3573 | hCoV-19/USA/MN-MDH-1216/2020\|EPI_ISL_477305\|2020-0 | ···················· | ···················· | ····················· | ···················· | ···················· | ···················· | |
| D | 3574 | hCoV-19/USA/MN-MDH-1217/2020\|EPI_ISL_477306\|2020-0 | ···················· | ···················· | ····················· | ···················· | ···················· | ···················· | |
| D | 3575 | hCoV-19/USA/MN-MDH-1221/2020\|EPI_ISL_477310\|2020-0 | ···················· | ···················· | ····················· | ···················· | ···················· | ···················· | |
| D | 3576 | hCoV-19/USA/MN-MDH-1222/2020\|EPI_ISL_477311\|2020-0 | ···················· | ···················· | ····················· | ···················· | ···················· | ···················· | |
| D | 3577 | hCoV-19/USA/MN-MDH-1223/2020\|EPI_ISL_477312\|2020-0 | ···················· | ···················· | ····················· | ···················· | ···················· | ···················· | |
| D | 3578 | hCoV-19/England/NORT-294B95/2020\|EPI_ISL_478608\|20 | ···················· | ···················· | ····················· | ···················· | ···················· | ···················· | |
| D | 3579 | hCoV-19/England/NORT-29545D/2020\|EPI_ISL_478636\|20 | ···················· | ···················· | ····················· | ···················· | ···················· | ···················· | |
| D | 3580 | hCoV-19/England/NORT-29546C/2020\|EPI_ISL_478637\|20 | ···················· | ···················· | ····················· | ···················· | ···················· | ···················· | |
| D | 3581 | hCoV-19/England/NORT-2959F4/2020\|EPI_ISL_478645\|20 | ···················· | ···················· | ····················· | ···················· | ···················· | ···················· | |
| D | 3582 | hCoV-19/England/NORT-295AA6/2020\|EPI_ISL_478648\|20 | ···················· | ···················· | ····················· | ···················· | ···················· | ···················· | |
| D | 3583 | hCoV-19/England/NORT-295B76/2020\|EPI_ISL_478652\|20 | ···················· | ·A·················· | ····················· | ···················· | ···················· | ···················· | |
| D | 3584 | hCoV-19/England/NORT-295BD0/2020\|EPI_ISL_478655\|20 | ···················· | ···················· | ····················· | ···················· | ···················· | ···················· | |
| D | 3585 | hCoV-19/England/20119000304/2020\|EPI_ISL_465001\|20 | ···················· | ···················· | ····················· | ···················· | ···················· | ···················· | |
| D | 3586 | hCoV-19/England/20119000404/2020\|EPI_ISL_465002\|20 | ···················· | ···················· | ····················· | ···················· | ··N················· | ···················· | |
| D | 3589 | hCoV-19/England/20119009104/2020\|EPI_ISL_465024\|20 | ···················· | ···················· | ····················· | ···················· | ···················· | ···················· | |
| D | 3590 | hCoV-19/England/20170003404/2020\|EPI_ISL_465270\|20 | ···················· | ···················· | ····················· | ···················· | ···················· | ···················· | |
| D | 3591 | hCoV-19/England/20170003504/2020\|EPI_ISL_465271\|20 | ···················· | ···················· | ····················· | ···················· | ···················· | ···················· | |
| D | 3593 | hCoV-19/England/20170030604/2020\|EPI_ISL_465273\|20 | ···················· | ···················· | ····················· | ···················· | ···················· | ···················· | |
| D | 3594 | hCoV-19/England/20170030904/2020\|EPI_ISL_465274\|20 | ···················· | ···················· | ····················· | ···················· | ···················· | ···················· | |
| D | 3595 | hCoV-19/England/20170031904/2020\|EPI_ISL_465275\|20 | ···················· | ···················· | ····················· | ···················· | ···················· | ···················· | |
| D | 3596 | hCoV-19/England/20170032004/2020\|EPI_ISL_465276\|20 | ···················· | ···················· | ····················· | ···················· | ···················· | ···················· | |
| D | 3597 | hCoV-19/England/20170032104/2020\|EPI_ISL_465277\|20 | ···················· | ···················· | ····················· | ···················· | ···················· | ···················· | |
| D | 3598 | hCoV-19/England/20170032204/2020\|EPI_ISL_465278\|20 | ···················· | ···················· | ····················· | ···················· | ···················· | ···················· | |
| D | 3600 | hCoV-19/England/20170033804/2020\|EPI_ISL_465280\|20 | ···················· | ···················· | ····················· | ···················· | ···················· | ···················· | |
| D | 3603 | hCoV-19/England/20170035404/2020\|EPI_ISL_465283\|20 | ···················· | ···················· | ····················· | ···················· | ···················· | ···················· | |
| D | 3606 | hCoV-19/England/20170036904/2020\|EPI_ISL_465287\|20 | ···················· | ···················· | ····················· | ···················· | ···················· | ···················· | |
| D | 3607 | hCoV-19/England/20170037604/2020\|EPI_ISL_465289\|20 | ···················· | ···················· | ····················· | ···················· | ···················· | ···················· | |
| D | 3608 | hCoV-19/England/20170037804/2020\|EPI_ISL_465290\|20 | ···················· | ···················· | ····················· | ···················· | ···················· | ···················· | |
| D | 3609 | hCoV-19/England/20170042504/2020\|EPI_ISL_465294\|20 | ···················· | ···················· | ····················· | ···················· | ···················· | ···················· | |
| D | 3610 | hCoV-19/England/20170047404/2020\|EPI_ISL_465297\|20 | ···················· | ···················· | ····················· | ···················· | ···················· | ··N··········NN····· | 1 |
| D | 3611 | hCoV-19/England/20170047704/2020\|EPI_ISL_465298\|20 | ···················· | ···················· | ····················· | ···················· | ···················· | ···················· | |
| D | 3613 | hCoV-19/England/20156041604/2020\|EPI_ISL_466311\|20 | ···················· | ···················· | ····················· | ···················· | ···················· | ···················· | |
| D | 3614 | hCoV-19/England/20156043304/2020\|EPI_ISL_466317\|20 | ···················· | ···················· | ····················· | ···················· | ···················· | ···················· | |
| D | 3618 | hCoV-19/England/20156047404/2020\|EPI_ISL_466324\|20 | ···················· | ···················· | ····················· | ···················· | ···················· | ···················· | |
| D | 3619 | hCoV-19/England/20156047804/2020\|EPI_ISL_466325\|20 | ···················· | ···················· | ····················· | ···················· | ···················· | ···················· | |
| D | 3620 | hCoV-19/England/20156047904/2020\|EPI_ISL_466326\|20 | ···················· | ···················· | ····················· | ···················· | ···················· | ···················· | |
| D | 3622 | hCoV-19/England/20156048204/2020\|EPI_ISL_466328\|20 | ···················· | ···················· | ····················· | ···················· | ···················· | ···················· | |
| D | 3623 | hCoV-19/England/20156050304/2020\|EPI_ISL_466336\|20 | ···················· | ···················· | ····················· | ···················· | ···················· | ··A················· | |
| D | 3624 | hCoV-19/England/20156050804/2020\|EPI_ISL_466338\|20 | ···················· | ···················· | ····················· | ···················· | ···················· | ···················· | |
| D | 3626 | hCoV-19/England/20156052404/2020\|EPI_ISL_466342\|20 | ···················· | ···················· | ····················· | ···················· | ···················· | ···················· | |
| D | 3627 | hCoV-19/England/20156053104/2020\|EPI_ISL_466343\|20 | ···················· | ···················· | ····················· | ···················· | ···················· | ···················· | |
| D | 3628 | hCoV-19/England/201580017/2020\|EPI_ISL_466346\|2020 | ···················· | ···················· | ····················· | ···················· | ···················· | ···················· | |
| D | 3629 | hCoV-19/England/201580036/2020\|EPI_ISL_466350\|2020 | ···················· | ···················· | ····················· | ···················· | ···················· | ···················· | |
| D | 3630 | hCoV-19/England/201580037/2020\|EPI_ISL_466351\|2020 | ···················· | ···················· | ····················· | ···················· | ···················· | ···················· | |
| D | 3631 | hCoV-19/England/201580039/2020\|EPI_ISL_466352\|2020 | ···················· | ···················· | ····················· | ···················· | ···················· | ···················· | |
| D | 3632 | hCoV-19/England/201580159/2020\|EPI_ISL_466355\|2020 | ···················· | ···················· | ····················· | ···················· | ···················· | ···················· | |
| D | 3633 | hCoV-19/England/201580161/2020\|EPI_ISL_466356\|2020 | ···················· | ···················· | ····················· | ···················· | ···················· | ···················· | |
| D | 3634 | hCoV-19/USA/NM-UNM-00023/2020\|EPI_ISL_467604\|2020- | ···················· | ···················· | ····················· | ···················· | ···················· | ···················· | |
| D | 3635 | hCoV-19/USA/NM-UNM-00027/2020\|EPI_ISL_467608\|2020- | ···················· | ···················· | ····················· | ···················· | ···················· | ···················· | |
| D | 3636 | hCoV-19/USA/NM-UNM-00029/2020\|EPI_ISL_467610\|2020- | ···················· | ···················· | ····················· | ···················· | ···················· | ···················· | |
| D | 3637 | hCoV-19/USA/NM-UNM-00030/2020\|EPI_ISL_467611\|2020- | ···················· | ···················· | ····················· | ···················· | ···················· | ···················· | |
| D | 3638 | hCoV-19/USA/NM-UNM-00031/2020\|EPI_ISL_467612\|2020- | ···················· | ···················· | ····················· | ···················· | ···················· | ···················· | |
| D | 3639 | hCoV-19/USA/NM-UNM-00043/2020\|EPI_ISL_467624\|2020- | ···················· | ···················· | ····················· | ···················· | ···················· | ···················· | |
| D | 3640 | hCoV-19/USA/NM-UNM-00083/2020\|EPI_ISL_467630\|2020- | ···················· | ···················· | ····················· | ···················· | ···················· | ···················· | |
| D | 3641 | hCoV-19/USA/NM-UNM-00085/2020\|EPI_ISL_467632\|2020- | ···················· | ···················· | ····················· | ···················· | ···················· | ···················· | |
| D | 3642 | hCoV-19/USA/NM-UNM-00086/2020\|EPI_ISL_467633\|2020- | ···················· | ···················· | ····················· | ···················· | ···················· | ···················· | |
| D | 3643 | hCoV-19/USA/NM-UNM-00087/2020\|EPI_ISL_467634\|2020- | ···················· | ···················· | ····················· | ···················· | ···················· | ···················· | |
| D | 3644 | hCoV-19/USA/NM-UNM-00110/2020\|EPI_ISL_467644\|2020- | ···················· | ···················· | ····················· | ···················· | ···················· | ···················· | |
| D | 3645 | hCoV-19/USA/NM-UNM-00133/2020\|EPI_ISL_467651\|2020- | ···················· | ···················· | ····················· | ···················· | ···················· | ···················· | |
| D | 3646 | hCoV-19/USA/NM-UNM-00134/2020\|EPI_ISL_467652\|2020- | ···················· | ···················· | ····················· | ···················· | ···················· | ···················· | |
| D | 3647 | hCoV-19/USA/NM-UNM-00142/2020\|EPI_ISL_467653\|2020- | ···················· | ···················· | ········G············ | ···················· | ···················· | ···················· | |
| D | 3648 | hCoV-19/USA/NM-UNM-00143/2020\|EPI_ISL_467654\|2020- | ···················· | ···················· | ····················· | ···················· | ···················· | ···················· | |

| | | | | | | | | |
|---|---|---|---|---|---|---|---|---|
| D | 3649 hCoV-19/USA/NM-UNM-00166/2020\|EPI_ISL_467661\|2020- | ···················· | ···················· | ···················· | ···················· | ···················· | ···················· | |
| D | 3650 hCoV-19/USA/NM-UNM-00168/2020\|EPI_ISL_467663\|2020- | ···················· | ···················· | ···················· | ···················· | ···················· | ···················· | |
| D | 3653 hCoV-19/Spain/PV-IBV-2092/2020\|EPI_ISL_468934\|2020 | ···················· | ···················· | ···················· | ···················· | ···················· | ···················· | |
| D | 3659 hCoV-19/Spain/PV-IBV-2129/2020\|EPI_ISL_468940\|2020 | ···················· | ···················· | ···················· | ···················· | ··N················· | ···················· | |
| D | 3660 hCoV-19/Spain/PV-IBV-2145/2020\|EPI_ISL_468941\|2020 | ···················· | ···················· | ···················· | ···················· | ···················· | ···················· | |
| D | 3664 hCoV-19/Spain/PV-IBV-2105/2020\|EPI_ISL_468945\|2020 | ···················· | ···················· | ···················· | ···················· | ···················· | ···················· | |
| D | 3667 hCoV-19/Spain/PV-IBV-2170/2020\|EPI_ISL_468948\|2020 | ···················· | ···················· | ···················· | ···················· | ···················· | ···················· | |
| D | 3668 hCoV-19/Spain/PV-IBV-2084/2020\|EPI_ISL_468949\|2020 | ···················· | ···················· | ···················· | ···················· | ··N················· | ···················· | |
| D | 3678 hCoV-19/Spain/IB-IBV-2591/2020\|EPI_ISL_468966\|2020 | ···················· | ···················· | ···················· | ···················· | ···················· | ···················· | |
| D | 3682 hCoV-19/Spain/IB-IBV-2542/2020\|EPI_ISL_468975\|2020 | ···················· | ···················· | ···················· | ···················· | ···················· | ···················· | |
| D | 3685 hCoV-19/Spain/IB-IBV-2513/2020\|EPI_ISL_468979\|2020 | ···················· | ···················· | ···················· | ···················· | ···················· | ···················· | |
| D | 3695 hCoV-19/Spain/IB-IBV-2520/2020\|EPI_ISL_468991\|2020 | ···················· | ···················· | ···················· | ···················· | ···················· | ···················· | |
| D | 3701 hCoV-19/England/NOTT-111EE3/2020\|EPI_ISL_472422\|20 | ···················· | ···················· | ···················· | ···················· | ···················· | ···················· | |
| D | 3702 hCoV-19/England/NOTT-111F4A/2020\|EPI_ISL_472427\|20 | ···················· | ···················· | ···················· | ···················· | ···················· | ···················· | |
| D | 3703 hCoV-19/England/NOTT-111F59/2020\|EPI_ISL_472428\|20 | ···················· | ···················· | ···················· | ···················· | ···················· | ···················· | |
| D | 3704 hCoV-19/England/NOTT-111F68/2020\|EPI_ISL_472429\|20 | ···················· | ···················· | ···················· | ···················· | ···················· | ···················· | |
| D | 3705 hCoV-19/England/CAMB-1B493C/2020\|EPI_ISL_473487\|20 | ···················· | ···················· | ···················· | ···················· | ···················· | ···················· | |
| D | 3706 hCoV-19/England/CAMB-1B494B/2020\|EPI_ISL_473488\|20 | ···················· | ···················· | ···················· | ···················· | ···················· | ···················· | |
| D | 3707 hCoV-19/England/CAMB-1B495A/2020\|EPI_ISL_473489\|20 | ···················· | ···················· | ···················· | ···················· | ···················· | ···················· | |
| D | 3709 hCoV-19/Scotland/CVR3572/2020\|EPI_ISL_473705\|2020- | ···················· | ···················· | ···················· | ···················· | ···················· | ···················· | |
| D | 3710 hCoV-19/Scotland/CVR3573/2020\|EPI_ISL_473706\|2020- | ···················· | ···················A | ···················· | ···················· | ···················· | ···················· | 1 |
| D | 3712 hCoV-19/Scotland/CVR3577/2020\|EPI_ISL_473709\|2020- | ···················· | ···················· | ···················· | ···················· | ···················· | ···················· | |
| D | 3714 hCoV-19/Scotland/CVR3582/2020\|EPI_ISL_473713\|2020- | ···················· | ···················· | ···················· | ···················· | ···················· | ···················· | |
| D | 3715 hCoV-19/Scotland/CVR3584/2020\|EPI_ISL_473715\|2020- | ···················· | ···················· | ···················· | ···················· | ···················· | ···················· | |
| D | 3716 hCoV-19/Scotland/CVR3586/2020\|EPI_ISL_473717\|2020- | ···················· | ···················A | ···················· | ···················· | ···················· | ···················· | 1 |
| D | 3718 hCoV-19/Scotland/CVR3672/2020\|EPI_ISL_473753\|2020- | ···················· | ···················· | ···················· | ···················· | ···················· | ···················· | |
| D | 3719 hCoV-19/Belgium/rega-0504484/2020\|EPI_ISL_477000\|2 | ···················· | ···················· | ···················· | ···················· | ···················· | ···················· | |
| D | 3720 hCoV-19/Belgium/rega-0505428/2020\|EPI_ISL_477001\|2 | ···················· | ···················· | ···················· | ···················· | ···················· | ···················· | |
| D | 3721 hCoV-19/Belgium/rega-0505485/2020\|EPI_ISL_477002\|2 | ···················· | ···················· | ···················· | ···················· | ···················· | ···················· | |
| D | 3722 hCoV-19/Belgium/rega-0505486/2020\|EPI_ISL_477003\|2 | ···················· | ···················· | ···················· | ···················· | ···················· | ···················· | |
| D | 3723 hCoV-19/Belgium/rega-0506487/2020\|EPI_ISL_477004\|2 | ···················· | ···················· | ···················· | ···················· | ···················· | ···················· | |
| D | 3724 hCoV-19/Belgium/rega-0507488/2020\|EPI_ISL_477005\|2 | ···················· | ···················· | ···················· | ···················· | ···················· | ···················· | |
| D | 3725 hCoV-19/India/KA-InStem-NCBS-0049/2020\|EPI_ISL_477 | ···················· | ···················· | ···················· | ···················· | ···················· | ···················· | |
| D | 3726 hCoV-19/India/KA-InStem-NCBS-0050/2020\|EPI_ISL_477 | ···················· | ···················· | ···················· | ···················· | ···················· | ···················· | |
| D | 3727 hCoV-19/India/KA-InStem-NCBS-0055/2020\|EPI_ISL_477 | ···················· | ·A·················· | ···················· | ···················· | ···················· | ···A················ | |
| D | 3728 hCoV-19/India/KA-InStem-NCBS-0061/2020\|EPI_ISL_477 | ···················· | ···················· | ···················· | ···················· | ···················· | ···················· | |
| D | 3729 hCoV-19/USA/MN-MDH-1182/2020\|EPI_ISL_477272\|2020-0 | ···················· | ·A·················· | ···················· | ···················· | ···················· | ···················· | |
| D | 3730 hCoV-19/USA/MN-MDH-1183/2020\|EPI_ISL_477273\|2020-0 | ···················· | ·A·················· | ···················· | ···················· | ···················· | ···················· | |
| D | 3731 hCoV-19/USA/MN-MDH-1184/2020\|EPI_ISL_477274\|2020-0 | ···················· | ·A·················· | ···················· | ···················· | ···················· | ···················· | |
| D | 3732 hCoV-19/USA/MN-MDH-1185/2020\|EPI_ISL_477275\|2020-0 | ···················· | ·A·················· | ···················· | ···················· | ···················· | ···················· | |
| D | 3733 hCoV-19/USA/MN-MDH-1186/2020\|EPI_ISL_477276\|2020-0 | ···················· | ···················· | ···················· | ···················· | ···················· | ···················· | |
| D | 3734 hCoV-19/USA/MN-MDH-1187/2020\|EPI_ISL_477277\|2020-0 | ···················· | ·A·················· | ···················· | ···················· | ···················· | ···················· | |
| D | 3735 hCoV-19/USA/MN-MDH-1188/2020\|EPI_ISL_477278\|2020-0 | ···················· | ·A·················· | ···················· | ···················· | ···················· | ···················· | |
| D | 3736 hCoV-19/USA/MN-MDH-1189/2020\|EPI_ISL_477279\|2020-0 | ···················· | ·A·················· | ···················· | ···················· | ···················· | ···················· | |
| D | 3737 hCoV-19/USA/MN-MDH-1190/2020\|EPI_ISL_477280\|2020-0 | ···················· | ·A·················· | ···················· | ···················· | ···················· | ···················· | |
| D | 3738 hCoV-19/USA/MN-MDH-1191/2020\|EPI_ISL_477281\|2020-0 | ···················· | ·A·················· | ···················· | ···················· | ···················· | ···················· | |
| D | 3739 hCoV-19/USA/MN-MDH-1192/2020\|EPI_ISL_477282\|2020-0 | ···················· | ·A·················· | ···················· | ···················· | ···················· | ···················· | |
| D | 3740 hCoV-19/USA/MN-MDH-1193/2020\|EPI_ISL_477283\|2020-0 | ···················· | ·A·················· | ···················· | ···················· | ···················· | ···················· | |
| D | 3741 hCoV-19/USA/MN-MDH-1194/2020\|EPI_ISL_477284\|2020-0 | ···················· | ·A·················· | ···················· | ···················· | ···················· | ···················· | |
| D | 3742 hCoV-19/USA/MN-MDH-1195/2020\|EPI_ISL_477285\|2020-0 | ···················· | ·A·················· | ···················· | ···················· | ···················· | ···················· | |
| D | 3743 hCoV-19/USA/MN-MDH-1196/2020\|EPI_ISL_477286\|2020-0 | ···················· | ·A·················· | ···················· | ···················· | ···················· | ···················· | |
| D | 3744 hCoV-19/USA/MN-MDH-1197/2020\|EPI_ISL_477287\|2020-0 | ···················· | ·A·················· | ···················· | ···················· | ···················· | ···················· | |
| D | 3745 hCoV-19/USA/MN-MDH-1198/2020\|EPI_ISL_477288\|2020-0 | ···················· | ·A·················· | ···················· | ···················· | ···················· | ···················· | |
| D | 3746 hCoV-19/USA/MN-MDH-1199/2020\|EPI_ISL_477289\|2020-0 | ···················· | ·A·················· | ···················· | ···················· | ···················· | ···················· | |
| D | 3747 hCoV-19/USA/MN-MDH-1201/2020\|EPI_ISL_477290\|2020-0 | ···················· | ·A·················· | ···················· | ···················· | ···················· | ···················· | |
| D | 3748 hCoV-19/USA/MN-MDH-1202/2020\|EPI_ISL_477291\|2020-0 | ···················· | ···················· | ···················· | ···················· | ···················· | ···················· | |
| D | 3749 hCoV-19/USA/MN-MDH-1203/2020\|EPI_ISL_477292\|2020-0 | ···················· | ···················· | ···················· | ···················· | ···················· | ···················· | |
| D | 3750 hCoV-19/USA/MN-MDH-1204/2020\|EPI_ISL_477293\|2020-0 | ···················· | ···················· | ···················· | ···················· | ···················· | ···················· | |
| D | 3751 hCoV-19/USA/MN-MDH-1205/2020\|EPI_ISL_477294\|2020-0 | ···················· | ···················· | ···················· | ···················· | ···················· | ···················· | |
| D | 3752 hCoV-19/USA/MN-MDH-1206/2020\|EPI_ISL_477295\|2020-0 | ···················· | ···················· | ···················· | ···················· | ···················· | ···················· | |
| D | 3753 hCoV-19/USA/MN-MDH-1207/2020\|EPI_ISL_477296\|2020-0 | ···················· | ···················· | ···················· | ···················· | ···················· | ···················· | |
| D | 3754 hCoV-19/USA/MN-MDH-1208/2020\|EPI_ISL_477297\|2020-0 | ···················· | ···················· | ···················· | ···················· | ···················· | ···················· | |
| D | 3755 hCoV-19/USA/MN-MDH-1209/2020\|EPI_ISL_477298\|2020-0 | ···················· | ···················· | ···················· | ···················· | ···················· | ···················· | |
| D | 3756 hCoV-19/USA/MN-MDH-1210/2020\|EPI_ISL_477299\|2020-0 | ···················· | ···················· | ···················· | ···················· | ···················· | ···················· | |
| D | 3757 hCoV-19/England/LCST-24D684D/2020\|EPI_ISL_478301\|2 | ···················· | ···················· | ···················· | ···················· | ···················· | ···················· | |
| D | 3758 hCoV-19/England/LCST-24D687A/2020\|EPI_ISL_478304\|2 | ···················· | ···················· | ···················· | ···················· | ···················· | ···················· | |
| D | 3759 hCoV-19/England/LCST-24D6889/2020\|EPI_ISL_478305\|2 | ···················· | ···················· | ···················· | ···················· | ···················· | ···················· | |
| D | 3760 hCoV-19/England/LCST-24D6898/2020\|EPI_ISL_478306\|2 | ···················· | ···················· | ···················· | ···················· | ···················· | ···················· | |
| D | 3761 hCoV-19/England/LCST-24D68A7/2020\|EPI_ISL_478307\|2 | ···················· | ···················· | ···················· | ···················· | ···················· | ···················· | |
| D | 3762 hCoV-19/England/LCST-24D68F2/2020\|EPI_ISL_478311\|2 | ···················· | ···················· | ···················· | ···················· | ···················· | ···················· | |
| D | 3763 hCoV-19/England/LCST-24D692C/2020\|EPI_ISL_478312\|2 | ···················· | ···················· | ···················· | ···················· | ···················· | ···················· | |
| D | 3764 hCoV-19/England/LCST-24D69B3/2020\|EPI_ISL_478315\|2 | ···················· | ···················· | ···················· | ···················· | ···················· | ···················· | |
| D | 3765 hCoV-19/England/LCST-24D69C2/2020\|EPI_ISL_478316\|2 | ···················· | ···················· | ···················· | ···················· | ···················· | ···················· | |
| D | 3766 hCoV-19/England/LCST-24D69D1/2020\|EPI_ISL_478317\|2 | ···················· | ···················· | ···················· | ···················· | ···················· | ···················· | |
| D | 3767 hCoV-19/England/LCST-24D6A29/2020\|EPI_ISL_478320\|2 | ···················· | ···················· | ···················· | ···················· | ···················· | ···················· | |
| D | 3768 hCoV-19/England/LCST-24D6A38/2020\|EPI_ISL_478321\|2 | ···················· | ···················· | ···················· | ···················· | ···················· | ···················· | |
| D | 3769 hCoV-19/England/LCST-24D6A74/2020\|EPI_ISL_478324\|2 | ···················· | ···················· | ···················· | ···················· | ···················· | ···················· | |
| D | 3770 hCoV-19/England/LCST-24D6A83/2020\|EPI_ISL_478325\|2 | ···················· | ···················· | ···················· | ···················· | ···················· | ···················· | |

| | | | | | | | | |
|---|---|---|---|---|---|---|---|---|
| D | 3771 | hCoV-19/England/LCST-24D6AA1/2020\|EPI_ISL_478327\|2 | .................... | .................... | .................... | .................... | .................... | .................... |
| D | 3772 | hCoV-19/England/LCST-24D6AB0/2020\|EPI_ISL_478328\|2 | .................... | .................... | .................... | .................... | .................... | .................... |
| D | 3773 | hCoV-19/England/LCST-24D6ACF/2020\|EPI_ISL_478329\|2 | .................... | .................... | .................... | .................... | .................... | .................... |
| D | 3774 | hCoV-19/England/LCST-24D6B26/2020\|EPI_ISL_478330\|2 | .................... | .................... | .................... | .................... | .................... | .................... |
| D | 3775 | hCoV-19/England/LCST-24D6B35/2020\|EPI_ISL_478331\|2 | .................... | .................... | .................... | .................... | .................... | .................... |
| D | 3776 | hCoV-19/England/LCST-24D6B62/2020\|EPI_ISL_478334\|2 | .................... | .................... | .................... | .................... | .................... | .................... |
| D | 3777 | hCoV-19/England/LCST-24D6B71/2020\|EPI_ISL_478335\|2 | .................... | .................... | .................... | .................... | .................... | .................... |
| D | 3778 | hCoV-19/England/LCST-24D6B80/2020\|EPI_ISL_478336\|2 | .................... | .................... | .................... | .................... | .................... | .................... |
| D | 3779 | hCoV-19/England/NORT-284B5C/2020\|EPI_ISL_478579\|20 | .................... | .................... | .................... | .................... | .................... | .................... |
| D | 3780 | hCoV-19/England/NORT-284B98/2020\|EPI_ISL_478580\|20 | .................... | .................... | .................... | .................... | .................... | .................... |
| D | 3781 | hCoV-19/England/NORT-284BA7/2020\|EPI_ISL_478581\|20 | .................... | .................... | .................... | .................... | .................... | .................... |
| D | 3782 | hCoV-19/England/NORT-284BB6/2020\|EPI_ISL_478582\|20 | .................... | .................... | .................... | .................... | .................... | .................... |
| D | 3783 | hCoV-19/England/NORT-284BC5/2020\|EPI_ISL_478583\|20 | .................... | .................... | .................... | .................... | .................... | .................... |
| D | 3784 | hCoV-19/England/NORT-284BD4/2020\|EPI_ISL_478584\|20 | .................... | .................... | .................... | .................... | .................... | .................... |
| D | 3785 | hCoV-19/England/NORT-284C3B/2020\|EPI_ISL_478585\|20 | .................... | .................... | .................... | .................... | .................... | .................... |
| D | 3786 | hCoV-19/England/NORT-284C86/2020\|EPI_ISL_478586\|20 | .................... | .................... | .................... | .................... | .................... | .................... |
| D | 3787 | hCoV-19/England/NORT-284C95/2020\|EPI_ISL_478587\|20 | .................... | .................... | .................... | .................... | .................... | .................... |
| D | 3788 | hCoV-19/England/NORT-284CA4/2020\|EPI_ISL_478588\|20 | .................... | .................... | .................... | .................... | .................... | .................... |
| D | 3789 | hCoV-19/England/NORT-284D74/2020\|EPI_ISL_478591\|20 | .................... | .................... | .................... | .................... | .................... | .................... |
| D | 3790 | hCoV-19/England/NORT-284D83/2020\|EPI_ISL_478592\|20 | .................... | .................... | .................... | .................... | .................... | .................... |
| D | 3791 | hCoV-19/England/NORT-284D92/2020\|EPI_ISL_478593\|20 | .................... | .................... | .................... | .................... | .................... | .................... |
| D | 3792 | hCoV-19/England/NORT-284DB0/2020\|EPI_ISL_478594\|20 | .................... | .................... | .................... | .................... | .................... | .................... |
| D | 3793 | hCoV-19/England/NORT-284E80/2020\|EPI_ISL_478596\|20 | .................... | .................... | .................... | .................... | .................... | .................... |
| D | 3794 | hCoV-19/Singapore/522/2020\|EPI_ISL_479600\|2020-04- | .................... | .................... | .................... | .................... | .................... | .................... |
| D | 3795 | hCoV-19/Singapore/523/2020\|EPI_ISL_479601\|2020-04- | .................... | .................... | .................... | .................... | .................... | .................... |
| D | 3796 | hCoV-19/Singapore/524/2020\|EPI_ISL_479602\|2020-04- | .................... | .................... | .................... | .................... | .................... | .................... |
| D | 3797 | hCoV-19/Singapore/525/2020\|EPI_ISL_479603\|2020-04- | .................... | .................... | .................... | .................... | .................... | .................... |
| D | 3798 | hCoV-19/Italy/FVG-ICGEB-bal1A/2020\|EPI_ISL_479616\| | .................... | .................... | .................... | .................... | .................... | .................... |
| D | 3799 | hCoV-19/Italy/FVG-ICGEB-bal1B/2020\|EPI_ISL_479617\| | .................... | .................... | .................... | .................... | .................... | .................... |
| D | 3800 | hCoV-19/Italy/FVG-ICGEB-bal16/2020\|EPI_ISL_479618\| | .................... | .................... | .................... | .................... | .................... | .................... |
| D | 3801 | hCoV-19/Italy/FVG-ICGEB-atr18/2020\|EPI_ISL_479619\| | .................... | .................... | .................... | .................... | .................... | .................... |
| D | 3802 | hCoV-19/Russia/CRIE182061/2020\|EPI_ISL_479620\|2020 | .................... | .................... | .................... | .................... | .................... | .................... |
| D | 3803 | hCoV-19/Russia/CRIE161454/2020\|EPI_ISL_479621\|2020 | .................... | .................... | .................... | .................... | .................... | .................... |
| D | 3804 | hCoV-19/Russia/CRIE164776/2020\|EPI_ISL_479622\|2020 | .................... | .................... | .................... | .................... | .................... | .................... |
| D | 3805 | hCoV-19/Russia/CRIE180117/2020\|EPI_ISL_479623\|2020 | .................... | .................... | .................... | .................... | .................... | .................... |
| D | 3806 | hCoV-19/Russia/CRIE155506/2020\|EPI_ISL_479624\|2020 | .................... | .................... | .................... | .................... | .................... | .................... |
| D | 3807 | hCoV-19/USA/NY-NYU-VC-008/2020\|EPI_ISL_479664\|2020 | .................... | .................... | .................... | .................... | .................... | .................... |
| D | 3808 | hCoV-19/USA/NY-NYU-VC-009a/2020\|EPI_ISL_479665\|202 | .................... | .................... | .................... | .................... | .................... | .................... |
| D | 3809 | hCoV-19/USA/NY-NYU-VC-010/2020\|EPI_ISL_479667\|2020 | .................... | .................... | .................... | .................... | .................... | .................... |
| D | 3810 | hCoV-19/Bangladesh/DNAS-CPH-isl-441/2020\|EPI_ISL_6 | .................... | .................... | .................... | .................... | .................... | .................... |
| D | 3811 | hCoV-19/Bangladesh/DNAS-CPH-isl-475/2020\|EPI_ISL_6 | .................... | .................... | .................... | .................... | .................... | .................... |
| D | 3812 | hCoV-19/Bangladesh/DNAS-CPH-isl-428/2020\|EPI_ISL_6 | .................... | .................... | .................... | .................... | .................... | .................... |
| D | 3813 | hCoV-19/Bangladesh/DNAS-CPH-isl-521/2020\|EPI_ISL_6 | .................... | .................... | .................... | .................... | .................... | .................... |
| D | 3814 | hCoV-19/Bangladesh/DNAS-P-isl-T/2020\|EPI_ISL_60588 | .................... | .................... | .................... | .................... | .................... | .................... |
| D | 3815 | hCoV-19/Bangladesh/DNAS-CPH-isl-417/2020\|EPI_ISL_6 | .................... | .................... | .................... | .................... | .................... | .................... |
| D | 3816 | hCoV-19/Bangladesh/DNAS-CPH-isl-485/2020\|EPI_ISL_6 | .................... | .................... | .................... | .................... | .................... | .................... |
| D | 3817 | hCoV-19/Bangladesh/DNAS-CPH-isl-509/2020\|EPI_ISL_6 | .................... | .................... | .................... | .................... | .................... | .................... |
| D | 3818 | hCoV-19/Bangladesh/DNAS-RAB-isl-86/2020\|EPI_ISL_60 | .................... | .................... | .................... | .................... | .................... | .................... |
| D | 3819 | hCoV-19/Bangladesh/DNAS-CPH-isl-426/2020\|EPI_ISL_6 | .................... | .................... | .................... | .................... | .................... | .................... |
| D | 3820 | hCoV-19/Bangladesh/DNAS-CPH-isl-461/2020\|EPI_ISL_6 | .................... | .................... | .................... | .................... | .................... | .................... |
| D | 3821 | hCoV-19/Bangladesh/DNAS-CPH-isl-437/2020\|EPI_ISL_6 | .................... | .................... | .................... | .................... | .................... | .................... |
| D | 3822 | hCoV-19/Bangladesh/DNAS-RAB-isl-114/2020\|EPI_ISL_6 | .................... | .................... | .................... | .................... | .................... | .................... |
| D | 3823 | hCoV-19/Bangladesh/DNAS-RAB-isl-118/2020\|EPI_ISL_6 | .................... | .................... | .................... | .................... | .................... | .................... |
| D | 3824 | hCoV-19/Bangladesh/DNAS-CPH-isl-537/2020\|EPI_ISL_6 | .................... | .................... | .................... | .................... | .................... | .................... |
| D | 3825 | hCoV-19/Bangladesh/DNAS-BSH-isl-9/2020\|EPI_ISL_605 | .................... | .................... | .................... | .................... | .................... | .................... |
| D | 3826 | hCoV-19/Bangladesh/DNAS-isl-113/2020\|EPI_ISL_60590 | .................... | .................... | .................... | .................... | .................... | .................... |
| D | 3827 | hCoV-19/Bangladesh/DNAS-isl-1-003/2020\|EPI_ISL_605 | .................... | .................... | .................... | .................... | .................... | .................... |
| D | 3828 | hCoV-19/Bangladesh/DNAS-isl-75/2020\|EPI_ISL_605911 | .................... | .................... | .................... | .................... | .................... | .................... |
| D | 3829 | hCoV-19/Bangladesh/DNAS-isl-95/2020\|EPI_ISL_605912 | .................... | .................... | .................... | .................... | .................... | .................... |
| D | 3830 | hCoV-19/Bangladesh/DNAS-isl-116/2020\|EPI_ISL_60591 | .................... | .................... | .................... | .................... | .................... | .................... |
| D | 3831 | hCoV-19/Bangladesh/DNAS-isl-6299/2020\|EPI_ISL_6059 | .................... | .................... | .................... | .................... | .................... | .................... |
| D | 3832 | hCoV-19/Bangladesh/DNAS-isl-6840/2020\|EPI_ISL_6059 | .................... | .................... | .................... | .................... | .................... | .................... |
| D | 3833 | hCoV-19/Bangladesh/DNAS-isl-6841/2020\|EPI_ISL_6059 | .................... | .................... | .................... | .................... | .................... | .................... |
| D | 3834 | hCoV-19/Bangladesh/DNAS-isl-8225/2020\|EPI_ISL_6059 | .................... | .................... | .................... | .................... | .................... | ...............A.... |
| D | 3835 | hCoV-19/Bangladesh/DNAS-isl-6300/2020\|EPI_ISL_6059 | .................... | .................... | .................... | .................... | .................... | .................... |
| D | 3836 | hCoV-19/Bangladesh/DNAS-isl-2/2020\|EPI_ISL_605919\| | .................... | .................... | .................... | .................... | .................... | .................... |
| D | 3837 | hCoV-19/Bangladesh/DNAS-isl-6302/2020\|EPI_ISL_6059 | .................... | .................... | .................... | .................... | .................... | .................... |
| D | 3838 | hCoV-19/Bangladesh/DNAS-isl-17/2020\|EPI_ISL_605921 | .................... | .................... | .................... | .................... | .................... | .................... |
| D | 3839 | hCoV-19/Bangladesh/DNAS-isl-6301/2020\|EPI_ISL_6059 | .................... | .................... | .................... | .................... | .................... | .................... |
| D | 3840 | hCoV-19/Bangladesh/DNAS-isl-4/2020\|EPI_ISL_605923\| | .................... | .................... | .................... | .................... | .................... | .................... |
| D | 3841 | hCoV-19/USA/MI-MDHHS-SC20618/2020\|EPI_ISL_462700\|2 | .................... | .................... | .................... | .................... | .................... | .................... |
| D | 3842 | hCoV-19/USA/MI-MDHHS-SC20619/2020\|EPI_ISL_462701\|2 | .................... | .................... | .................... | .................... | .................... | .................... |
| D | 3843 | hCoV-19/USA/MI-MDHHS-SC20625/2020\|EPI_ISL_462707\|2 | .................... | .................... | .................... | .................... | .................... | .................... |
| D | 3844 | hCoV-19/USA/MI-MDHHS-SC20626/2020\|EPI_ISL_462708\|2 | .................... | .................... | .................... | .................... | .................... | .................... |
| D | 3845 | hCoV-19/USA/MI-MDHHS-SC20637/2020\|EPI_ISL_462719\|2 | .................... | .................... | .................... | .................... | .................... | .................... |
| D | 3846 | hCoV-19/USA/MI-MDHHS-SC20638/2020\|EPI_ISL_462720\|2 | .................... | .................... | .................... | .................... | .................... | .................... |
| D | 3847 | hCoV-19/USA/MI-MDHHS-SC20643/2020\|EPI_ISL_462725\|2 | .................... | .................... | .................... | .................... | .................... | .................... |
| D | 3848 | hCoV-19/USA/MI-MDHHS-SC20649/2020\|EPI_ISL_462731\|2 | .................... | .................... | .................... | .................... | .................... | .................... |

| | | | | | | | | | |
|---|---|---|---|---|---|---|---|---|---|
| D | 3849 | hCoV-19/USA/MI-MDHHS-SC20650/2020\|EPI_ISL_462732\|2 | ···················· | ···················· | ···················· | ···················· | ···················· | ···················· | |
| D | 3850 | hCoV-19/USA/MI-MDHHS-SC20651/2020\|EPI_ISL_462733\|2 | ···················· | ···················· | ···················· | ···················· | ···················· | ···················· | |
| D | 3851 | hCoV-19/USA/MI-MDHHS-SC20652/2020\|EPI_ISL_462734\|2 | ···················· | ···················· | ···················· | ···················· | ···················· | ···················· | |
| D | 3852 | hCoV-19/USA/MI-MDHHS-SC20654/2020\|EPI_ISL_462736\|2 | ···················· | ···················· | ···················· | ···················· | ···················· | ···················· | |
| D | 3853 | hCoV-19/Canada/BC_18165111/2020\|EPI_ISL_462757\|202 | ···················· | ···················· | ···················· | ···················· | ···················· | ···················· | |
| D | 3854 | hCoV-19/Canada/BC_18507914/2020\|EPI_ISL_462758\|202 | ···················· | ···················· | ···················· | ···················· | ···················· | ···················· | |
| D | 3855 | hCoV-19/Canada/BC_18547866/2020\|EPI_ISL_462759\|202 | ···················· | ···················· | ···················· | ···················· | ···················· | ···················· | |
| D | 3856 | hCoV-19/Canada/BC_18969070/2020\|EPI_ISL_462760\|202 | ···················· | ···················· | ···················· | ···················· | ···················· | ···················· | |
| D | 3857 | hCoV-19/Canada/BC_19421714/2020\|EPI_ISL_462761\|202 | ···················· | ···················· | ···················· | ···················· | ···················· | ···················· | |
| D | 3858 | hCoV-19/Canada/BC_19518330/2020\|EPI_ISL_462765\|202 | ···················· | ···················· | ···················· | ···················· | ···················· | ···················· | |
| D | 3859 | hCoV-19/Canada/BC_19677629/2020\|EPI_ISL_462766\|202 | ···················· | ···················· | ···················· | ···················· | ···················· | ···················· | |
| D | 3860 | hCoV-19/Canada/BC_19703911/2020\|EPI_ISL_462767\|202 | ···················· | ···················· | ···················· | ···················· | ···················· | ···················· | |
| D | 3861 | hCoV-19/Canada/BC_19704814/2020\|EPI_ISL_462768\|202 | ···················· | ···················· | ···················· | ···················· | ···················· | ···················· | |
| D | 3862 | hCoV-19/Canada/BC_19785113/2020\|EPI_ISL_462769\|202 | ···················· | ···················· | ···················· | ···················· | ···················· | ···················· | |
| D | 3863 | hCoV-19/Canada/BC_20332712/2020\|EPI_ISL_462771\|202 | ···················· | ···················· | ···················· | ···················· | ···················· | ···················· | |
| D | 3864 | hCoV-19/Canada/BC_20363111/2020\|EPI_ISL_462772\|202 | ···················· | ···················· | ···················· | ···················· | ···················· | ···················· | |
| D | 3865 | hCoV-19/Canada/BC_20432317/2020\|EPI_ISL_462774\|202 | ···················· | ···················· | ···················· | ···················· | ···················· | ···················· | |
| D | 3866 | hCoV-19/Canada/BC_20505516/2020\|EPI_ISL_462775\|202 | ···················· | ···················· | ···················· | ···················· | ···················· | ···················· | |
| D | 3867 | hCoV-19/Canada/BC_20534610/2020\|EPI_ISL_462776\|202 | ···················· | ···················· | ···················· | ···················· | ···················· | ···················· | |
| D | 3868 | hCoV-19/Canada/BC_20639310/2020\|EPI_ISL_462777\|202 | ···················· | ···················· | ···················· | ···················· | ···················· | ···················· | |
| D | 3869 | hCoV-19/Canada/BC_20693717/2020\|EPI_ISL_462778\|202 | ···················· | ···················· | ···················· | ···················· | ···················· | ···················· | |
| D | 3870 | hCoV-19/Canada/BC_20722019/2020\|EPI_ISL_462779\|202 | ···················· | ···················· | ···················· | ···················· | ···················· | ···················· | |
| D | 3871 | hCoV-19/Canada/BC_20730616/2020\|EPI_ISL_462780\|202 | ···················· | ···················· | ···················· | ···················· | ···················· | ···················· | |
| D | 3872 | hCoV-19/Canada/BC_20734315/2020\|EPI_ISL_462781\|202 | ···················· | ···················· | ···················· | ···················· | ···················· | ···················· | |
| D | 3873 | hCoV-19/Canada/BC_20847524/2020\|EPI_ISL_462782\|202 | ···················· | ···················· | ···················· | ···················· | ···················· | ···················· | |
| D | 3874 | hCoV-19/Canada/BC_20855313/2020\|EPI_ISL_462783\|202 | ···················· | ···················· | ···················· | ···················· | ···················· | ···················· | |
| D | 3875 | hCoV-19/Canada/BC_21008811/2020\|EPI_ISL_462784\|202 | ···················· | ···················· | ···················· | ···················· | ···················· | ···················· | |
| D | 3876 | hCoV-19/Canada/BC_21016029/2020\|EPI_ISL_462786\|202 | ···················· | ···················· | ···················· | ···················· | ···················· | ···················· | |
| D | 3877 | hCoV-19/Canada/BC_21022514/2020\|EPI_ISL_462787\|202 | ···················· | ···················· | ···················· | ···················· | ···················· | ···················· | |
| D | 3878 | hCoV-19/Canada/BC_21085515/2020\|EPI_ISL_462789\|202 | ···················· | ···················· | ···················· | ···················· | ···················· | ···················· | |
| D | 3879 | hCoV-19/Canada/BC_21105219/2020\|EPI_ISL_462790\|202 | ···················· | ···················· | ···················· | ···················· | ···················· | ···················· | |
| D | 3880 | hCoV-19/Canada/BC_21110314/2020\|EPI_ISL_462791\|202 | ···················· | ···················· | ···················· | ···················· | ···················· | ···················· | |
| D | 3881 | hCoV-19/Canada/BC_21127214/2020\|EPI_ISL_462792\|202 | ···················· | ···················· | ···················· | ···················· | ···················· | ···················· | |
| D | 3882 | hCoV-19/Canada/BC_21127913/2020\|EPI_ISL_462793\|202 | ···················· | ···················· | ···················· | ···················· | ···················· | ···················· | |
| D | 3883 | hCoV-19/Canada/BC_21146111/2020\|EPI_ISL_462794\|202 | ···················· | ···················· | ···················· | ···················· | ···················· | ···················· | |
| D | 3884 | hCoV-19/Canada/BC_21187418/2020\|EPI_ISL_462798\|202 | ···················· | ···················· | ···················· | ···················· | ···················· | ···················· | |
| D | 3888 | hCoV-19/England/20152067804/2020\|EPI_ISL_466240\|20 | ···················· | ···················· | ···················· | ···················· | ···················· | ···················· | |
| D | 3890 | hCoV-19/England/20154000804/2020\|EPI_ISL_466242\|20 | ···················· | ···················· | ···················· | ···················· | ···················· | ·N·················· | |
| D | 3891 | hCoV-19/England/20154002504/2020\|EPI_ISL_466246\|20 | ···················· | ···················· | ···················· | ···················· | ···················· | ···················· | |
| D | 3892 | hCoV-19/England/20154002704/2020\|EPI_ISL_466248\|20 | ···················· | ···················· | ···················· | ···················· | ···················· | ···················· | |
| D | 3894 | hCoV-19/England/20154003804/2020\|EPI_ISL_466252\|20 | ···················· | ···················· | ···················· | ···················· | ···················· | ···················· | |
| D | 3895 | hCoV-19/England/20154003904/2020\|EPI_ISL_466253\|20 | ···················· | ···················· | ···················· | ···················· | ···················· | ···················· | |
| D | 3896 | hCoV-19/England/20154004204/2020\|EPI_ISL_466254\|20 | ···················· | ···················· | ···················· | ···················· | ···················· | ···················· | |
| D | 3897 | hCoV-19/England/20154004304/2020\|EPI_ISL_466255\|20 | ···················· | ···················· | ···················· | ···················· | ···················· | ···················· | |
| D | 3899 | hCoV-19/England/20154029904/2020\|EPI_ISL_466258\|20 | ···················· | ···················· | ···················· | ···················· | ···················· | ···················· | |
| D | 3900 | hCoV-19/England/20154074204/2020\|EPI_ISL_466259\|20 | ···················· | ···················· | ···················· | ···················· | ···················· | ···················· | |
| D | 3902 | hCoV-19/England/20154074804/2020\|EPI_ISL_466261\|20 | ···················· | ···················· | ···················· | ···················· | ···················· | ···················· | |
| D | 3904 | hCoV-19/England/20154082104/2020\|EPI_ISL_466267\|20 | ···················· | ···················· | ···················· | ···················· | ···················· | ···················· | |
| D | 3905 | hCoV-19/England/20154083504/2020\|EPI_ISL_466271\|20 | ···················· | ···················· | ···················· | ···················· | ···················· | ···················· | |
| D | 3906 | hCoV-19/England/20154083604/2020\|EPI_ISL_466272\|20 | ···················· | ···················· | ···················· | ···················· | ···················· | ···················· | |
| D | 3908 | hCoV-19/England/20154088104/2020\|EPI_ISL_466275\|20 | ···················· | ···················· | ···················· | ···················· | ···················· | ···················· | |
| D | 3909 | hCoV-19/England/20156000204/2020\|EPI_ISL_466278\|20 | ···················· | ···················· | ···················· | ···················· | ···················· | ···················· | |
| D | 3910 | hCoV-19/USA/NM-UNM-00151/2020\|EPI_ISL_467572\|2020- | ···················· | ···················· | ···················· | ···················· | ···················· | ···················· | |
| D | 3911 | hCoV-19/USA/NM-UNM-00162/2020\|EPI_ISL_467577\|2020- | ···················· | ···················· | ···················· | ···················· | ···················· | ···················· | |
| D | 3912 | hCoV-19/USA/NM-UNM-00174/2020\|EPI_ISL_467583\|2020- | ···················· | ···················· | ···················· | ···················· | ···················· | ···················· | |
| D | 3913 | hCoV-19/USA/NM-UNM-00007/2020\|EPI_ISL_467590\|2020- | ···················· | ···················· | ···················· | ···················· | ···················· | ···················· | |
| D | 3914 | hCoV-19/USA/NM-UNM-00009/2020\|EPI_ISL_467591\|2020- | ···················· | ···················· | ···················· | ···················· | ···················· | ···················· | |
| D | 3915 | hCoV-19/USA/NM-UNM-00012/2020\|EPI_ISL_467594\|2020- | ···················· | ···················· | ···················· | ···················· | ···················· | ···················· | |
| D | 3916 | hCoV-19/USA/NM-UNM-00014/2020\|EPI_ISL_467596\|2020- | ···················· | ···················· | ···················· | ···················· | ···················· | ···················· | |
| D | 3917 | hCoV-19/USA/NM-UNM-00016/2020\|EPI_ISL_467598\|2020- | ···················· | ···················· | ···················· | ···················· | ···················· | ···················· | |
| D | 3918 | hCoV-19/USA/NM-UNM-00018/2020\|EPI_ISL_467599\|2020- | ···················· | ···················· | ···················· | ···················· | ···················· | ···················· | |
| D | 3919 | hCoV-19/USA/NM-UNM-00019/2020\|EPI_ISL_467600\|2020- | ···················· | ···················· | ···················· | ···················· | ···················· | ···················· | |
| D | 3920 | hCoV-19/Spain/PV-IBV-2122/2020\|EPI_ISL_468899\|2020 | ···················· | ···················· | ···················· | ···················· | ···················· | ···················· | |
| D | 3924 | hCoV-19/Spain/PV-IBV-2112/2020\|EPI_ISL_468904\|2020 | ···················· | ···················· | ···················· | ···················· | ···················· | ···················· | |
| D | 3930 | hCoV-19/Spain/PV-IBV-2086/2020\|EPI_ISL_468910\|2020 | ···················· | ···················· | ···················· | ···················· | ···················· | ···················· | |
| D | 3932 | hCoV-19/Spain/PV-IBV-2139/2020\|EPI_ISL_468912\|2020 | ···················· | ···················· | ···················· | ···················· | ···················· | ···················· | |
| D | 3934 | hCoV-19/Italy/APU-UniMI-703/2020\|EPI_ISL_468914\|20 | ···················· | ···················· | ···················· | ···················· | ···················· | ···················· | |
| D | 3937 | hCoV-19/Spain/PV-IBV-2104/2020\|EPI_ISL_468917\|2020 | ···················· | ···················· | ···················· | ···················· | ···················· | ···················· | |
| D | 3941 | hCoV-19/Spain/PV-IBV-2166/2020\|EPI_ISL_468921\|2020 | ···················· | ···················· | ···················· | ···················· | ···················· | ···················· | |
| D | 3947 | hCoV-19/Spain/PV-IBV-2089/2020\|EPI_ISL_468927\|2020 | ···················· | ···················· | ···················· | ···················· | ···················· | ···················· | |
| D | 3952 | hCoV-19/England/LIVE-A02DF/2020\|EPI_ISL_472100\|202 | ···················· | ···················· | ···················· | ···················· | ···················· | ···················· | |
| D | 3953 | hCoV-19/England/LIVE-A0318/2020\|EPI_ISL_472102\|202 | ···················· | ···················· | ···················· | ···················· | ···················· | ···················· | |
| D | 3954 | hCoV-19/England/LIVE-A0363/2020\|EPI_ISL_472104\|202 | ···················· | ···················A | ···················· | ···················· | ···················· | ···················· | 1 |
| D | 3955 | hCoV-19/England/LIVE-A0372/2020\|EPI_ISL_472105\|202 | ···················· | ···················A | ···················· | ···················· | ···················· | ···················· | 1 |
| D | 3956 | hCoV-19/England/LIVE-A03AF/2020\|EPI_ISL_472106\|202 | ···················· | ···················· | ···················· | ···················· | ···················· | ···················· | |
| D | 3957 | hCoV-19/England/LIVE-A03BE/2020\|EPI_ISL_472107\|202 | ···················· | ···················· | ···················· | ···················· | ···················· | ···················· | |
| D | 3958 | hCoV-19/England/LIVE-A03DC/2020\|EPI_ISL_472109\|202 | ···················· | ···················· | ···················· | ···················· | ···················· | ···················· | |
| D | 3959 | hCoV-19/England/LIVE-A0460/2020\|EPI_ISL_472111\|202 | ···················· | ···················· | ···················· | ···················· | ···················· | ···················· | |

| | | | | | | | | | |
|---|---|---|---|---|---|---|---|---|---|
| D | 3960 | hCoV-19/England/LIVE-A049D/2020\|EPI_ISL_472113\|202 | ···················· | ···················· | ···················· | ···················· | ···················· | ···················· | |
| D | 3961 | hCoV-19/England/LIVE-A04AC/2020\|EPI_ISL_472114\|202 | ···················· | ···················· | ···················· | ···················· | ···················· | ···················· | |
| D | 3962 | hCoV-19/England/LIVE-A04D9/2020\|EPI_ISL_472115\|202 | ···················· | ···················· | ···················· | ···················· | ···················· | ···················· | |
| D | 3963 | hCoV-19/England/LIVE-A0503/2020\|EPI_ISL_472116\|202 | ···················· | ···················· | ···················· | ···················· | ···················· | ···················· | |
| D | 3964 | hCoV-19/England/LIVE-A0521/2020\|EPI_ISL_472118\|202 | ···················· | ···················· | ···················· | ···················· | ···················· | ···················· | |
| D | 3965 | hCoV-19/England/LIVE-A0530/2020\|EPI_ISL_472119\|202 | ···················· | ···················· | ···················· | ···················· | ···················· | ···················· | |
| D | 3966 | hCoV-19/England/LIVE-A0600/2020\|EPI_ISL_472121\|202 | ···················· | ···················· | ···················· | ···················· | ···················· | ···················· | |
| D | 3967 | hCoV-19/England/LIVE-A062E/2020\|EPI_ISL_472122\|202 | ···················· | ···················· | ···················· | ···················· | ···················· | ···················· | |
| D | 3968 | hCoV-19/England/LIVE-A065B/2020\|EPI_ISL_472124\|202 | ···················· | ···················· | ···················· | ···················· | ···················· | ···················· | |
| D | 3969 | hCoV-19/England/LIVE-A06D3/2020\|EPI_ISL_472126\|202 | ···················· | ···················· | ···················· | ···················· | ···················· | ···················· | |
| D | 3970 | hCoV-19/England/LIVE-A06E2/2020\|EPI_ISL_472127\|202 | ···················· | ···················· | ···················· | ···················· | ···················· | ···················· | |
| D | 3971 | hCoV-19/England/LIVE-A071C/2020\|EPI_ISL_472129\|202 | ···················· | ···················· | ···················· | ···················· | ···················· | ···················· | |
| D | 3973 | hCoV-19/England/LIVE-A0758/2020\|EPI_ISL_472132\|202 | ···················· | ···················· | ···················· | ···················· | ···················· | ···················· | |
| D | 3974 | hCoV-19/England/LIVE-A0767/2020\|EPI_ISL_472133\|202 | ···················· | ···················· | ···················· | ···················· | ···················· | ···················· | |
| D | 3975 | hCoV-19/England/LIVE-A0776/2020\|EPI_ISL_472134\|202 | ···················· | ···················· | ···················· | ···················· | ···················· | ···················· | |
| D | 3976 | hCoV-19/England/LIVE-A0785/2020\|EPI_ISL_472135\|202 | ···················· | ···················· | ···················· | ···················· | ···················· | ···················· | |
| D | 3982 | hCoV-19/England/BIRM-6043F/2020\|EPI_ISL_473410\|202 | ···················· | ···················· | ···················· | ···················· | ···················· | ···················· | |
| D | 3983 | hCoV-19/England/BIRM-6045D/2020\|EPI_ISL_473412\|202 | ···················· | ···················· | ···················· | ···················· | ···················· | ···················· | |
| D | 3984 | hCoV-19/England/BIRM-6047B/2020\|EPI_ISL_473414\|202 | ···················· | ···················· | ···················· | ···················· | ···················· | ···················· | |
| D | 3985 | hCoV-19/England/BIRM-6048A/2020\|EPI_ISL_473415\|202 | ···················· | ···················· | ···················· | ···················· | ···················· | ···················· | |
| D | 3986 | hCoV-19/England/BIRM-60499/2020\|EPI_ISL_473416\|202 | ···················· | ···················· | ···················· | ···················· | ···················· | ···················· | |
| D | 3988 | hCoV-19/England/BIRM-604D5/2020\|EPI_ISL_473419\|202 | ···················· | ···················· | ···················· | ···················· | ···················· | ···················· | |
| D | 3989 | hCoV-19/England/BIRM-6053C/2020\|EPI_ISL_473422\|202 | ···················· | ···················· | ···················· | ···················· | ···················· | ···················· | |
| D | 3990 | hCoV-19/England/BIRM-6054B/2020\|EPI_ISL_473423\|202 | ···················· | ···················· | ···················· | ···················· | ···················· | ···················· | |
| D | 3992 | hCoV-19/England/BIRM-60FD4/2020\|EPI_ISL_473427\|202 | ···················· | ···················· | ···················· | ···················· | ···················· | ···················· | |
| D | 3993 | hCoV-19/England/BIRM-60FF2/2020\|EPI_ISL_473429\|202 | ···················· | ···················· | ···················· | ···················· | ···················· | ···················· | |
| D | 3994 | hCoV-19/England/BIRM-61164/2020\|EPI_ISL_473434\|202 | ···················· | ···················· | ···················· | ···················· | ···················· | ···················· | |
| D | 3995 | hCoV-19/England/BIRM-61173/2020\|EPI_ISL_473435\|202 | ···················· | ···················· | ···················· | ···················· | ···················· | ···················· | |
| D | 3996 | hCoV-19/England/BIRM-61182/2020\|EPI_ISL_473436\|202 | ···················· | ···················· | ···················· | ···················· | ···················· | ···················· | |
| D | 3997 | hCoV-19/England/BIRM-611BF/2020\|EPI_ISL_473438\|202 | ···················· | ···················· | ···················· | ···················· | ···················· | ···················· | |
| D | 3998 | hCoV-19/England/BIRM-611EC/2020\|EPI_ISL_473441\|202 | ···················· | ···················· | ···················· | ···················· | ···················· | ···················· | |
| D | 3999 | hCoV-19/England/BIRM-611FB/2020\|EPI_ISL_473442\|202 | ···················· | ···················· | ···················· | ···················· | ···················· | ···················· | |
| D | 4000 | hCoV-19/England/BIRM-61207/2020\|EPI_ISL_473443\|202 | ···················· | ···················· | ···················· | ···················· | ···················· | ···················· | |
| D | 4002 | hCoV-19/England/BIRM-61225/2020\|EPI_ISL_473445\|202 | ···················· | ···················· | ···················· | ···················· | ···················· | ···················· | |
| D | 4003 | hCoV-19/England/BIRM-61243/2020\|EPI_ISL_473446\|202 | ···················· | ···················· | ···················· | ···················· | ···················· | ···················· | |
| D | 4006 | hCoV-19/England/BIRM-612AD/2020\|EPI_ISL_473450\|202 | ···················· | ···················· | ···················· | ···················· | ···················· | ···················· | |
| D | 4007 | hCoV-19/England/CAMB-1B3EF2/2020\|EPI_ISL_473454\|20 | ···················· | ···················· | ···················· | ···················· | ···················· | ···················· | |
| D | 4008 | hCoV-19/England/CAMB-1B3F1D/2020\|EPI_ISL_473455\|20 | ···················· | ···················· | ···················· | ···················· | ···················· | ···················· | |
| D | 4009 | hCoV-19/England/CAMB-1B4125/2020\|EPI_ISL_473459\|20 | ···················· | ···················· | ···················· | ···················· | ···················· | ···················· | |
| D | 4010 | hCoV-19/England/CAMB-1B4240/2020\|EPI_ISL_473463\|20 | ···················· | ···················· | ···················· | ···················· | ···················· | ···················· | |
| D | 4011 | hCoV-19/England/CAMB-1B426E/2020\|EPI_ISL_473464\|20 | ···················· | ···················· | ···················· | ···················· | ···················· | ···················· | |
| D | 4012 | hCoV-19/England/CAMB-1B42C8/2020\|EPI_ISL_473466\|20 | ···················· | ···················· | ···················· | ···················· | ···················· | ···················· | |
| D | 4013 | hCoV-19/England/CAMB-1B43E3/2020\|EPI_ISL_473471\|20 | ···················· | ···················· | ···················· | ···················· | ···················· | ···················· | |
| D | 4014 | hCoV-19/England/CAMB-1B442C/2020\|EPI_ISL_473472\|20 | ···················· | ···················· | ···················· | ···················· | ···················· | ···················· | |
| D | 4015 | hCoV-19/England/CAMB-1B44E0/2020\|EPI_ISL_473474\|20 | ···················· | ···················· | ···················· | ···················· | ···················· | ···················· | |
| D | 4016 | hCoV-19/England/CAMB-1B45A1/2020\|EPI_ISL_473477\|20 | ···················· | ···················· | ···················· | ···················· | ···················· | ···················· | |
| D | 4017 | hCoV-19/England/CAMB-1B4671/2020\|EPI_ISL_473479\|20 | ···················· | ···················· | ···················· | ···················· | ···················· | ···················· | |
| D | 4018 | hCoV-19/England/CAMB-1B4732/2020\|EPI_ISL_473481\|20 | ···················· | ···················· | ···················· | ···················· | ···················· | ···················· | |
| D | 4019 | hCoV-19/England/CAMB-1B477E/2020\|EPI_ISL_473482\|20 | ···················· | ···················· | ··········T········· | ···················· | ···················· | ···················· | |
| D | 4020 | hCoV-19/England/CAMB-1B479C/2020\|EPI_ISL_473483\|20 | ···················· | ···················· | ···················· | ···················· | ···················· | ···················· | |
| D | 4021 | hCoV-19/England/CAMB-1B48A8/2020\|EPI_ISL_473484\|20 | ···················· | ···················· | ···················· | ···················· | ···················· | ···················· | |
| D | 4022 | hCoV-19/England/CAMB-1B4969/2020\|EPI_ISL_473490\|20 | ···················· | ···················· | ···················· | ···················· | ···················· | ···················· | |
| D | 4023 | hCoV-19/England/CAMB-1B4A57/2020\|EPI_ISL_473491\|20 | ···················· | ···················· | ···················· | ···················· | ···················· | ···················· | |
| D | 4025 | hCoV-19/England/CAMB-1B4ADF/2020\|EPI_ISL_473493\|20 | ···················· | ···················· | ···················· | ···················· | ···················· | ···················· | |
| D | 4026 | hCoV-19/England/CAMB-1B4B09/2020\|EPI_ISL_473496\|20 | ···················· | ···················· | ···················· | ···················· | ···················· | ···················· | |
| D | 4027 | hCoV-19/England/CAMB-1B4B36/2020\|EPI_ISL_473498\|20 | ···················· | ···················· | ···················· | ···················· | ···················· | ···················· | |
| D | 4028 | hCoV-19/England/CAMB-1B4B45/2020\|EPI_ISL_473499\|20 | ···················· | ···················· | ···················· | ···················· | ···················· | ···················· | |
| D | 4029 | hCoV-19/Wales/PHWC-3671B/2020\|EPI_ISL_474704\|2020- | ···················· | ···················· | ···················· | ···················· | ···················· | ···················· | |
| D | 4030 | hCoV-19/Wales/PHWC-369AC/2020\|EPI_ISL_474740\|2020- | ···················· | ···················· | ···················· | ···················· | ···················· | ···················· | |
| D | 4031 | hCoV-19/Wales/PHWC-36A30/2020\|EPI_ISL_474748\|2020- | ···················· | ···················· | ···················· | ···················· | ···················· | ···················· | |
| D | 4032 | hCoV-19/Wales/PHWC-36AA9/2020\|EPI_ISL_474755\|2020- | ···················· | ···················· | ···················· | ···················· | ···················· | ···················· | |
| D | 4033 | hCoV-19/Wales/PHWC-36B79/2020\|EPI_ISL_474768\|2020- | ···················· | ···················· | ···················· | ···················· | ···················· | ···················· | |
| D | 4034 | hCoV-19/Wales/PHWC-36C2B/2020\|EPI_ISL_474779\|2020- | ···················· | ···················· | ···················· | ···················· | ···················· | ···················· | |
| D | 4035 | hCoV-19/Wales/PHWC-36C3A/2020\|EPI_ISL_474780\|2020- | ···················· | ···················· | ···················· | ···················· | ···················· | ···················· | |
| D | 4036 | hCoV-19/Wales/PHWC-36C85/2020\|EPI_ISL_474785\|2020- | ···················· | ···················· | ···················· | ···················· | ···················· | ···················· | |
| D | 4037 | hCoV-19/Wales/PHWC-36D19/2020\|EPI_ISL_474794\|2020- | ···················· | ···················· | ···················· | ···················· | ···················· | ···················· | |
| D | 4038 | hCoV-19/Wales/PHWC-36D28/2020\|EPI_ISL_474795\|2020- | ···················· | ···················· | ···················· | ···················· | ···················· | ···················· | |
| D | 4041 | hCoV-19/Scotland/EDB6094/2020\|EPI_ISL_478216\|2020- | ···················· | ···················A | ···················· | ···················· | ···················· | ···················· | 1 |
| D | 4042 | hCoV-19/Scotland/EDB6095/2020\|EPI_ISL_478217\|2020- | ···················· | ·A·················· | ···················· | ···················· | ···················· | ···················· | |
| D | 4043 | hCoV-19/Scotland/EDB6096/2020\|EPI_ISL_478218\|2020- | ···················· | ···················A | ···················· | ···················· | ···················· | ···················· | 1 |
| D | 4044 | hCoV-19/Scotland/EDB6103/2020\|EPI_ISL_478224\|2020- | ···················· | ···················· | ···················· | ···················· | ···················· | ···················· | |
| D | 4045 | hCoV-19/Scotland/EDB6104/2020\|EPI_ISL_478225\|2020- | ···················· | ···················· | ···················· | ···················· | ···················· | ···················· | |
| D | 4046 | hCoV-19/Scotland/EDB6108/2020\|EPI_ISL_478226\|2020- | ···················· | ···················A | ···················· | ···················· | ···················· | ···················· | 1 |
| D | 4047 | hCoV-19/Scotland/EDB6578/2020\|EPI_ISL_478260\|2020- | ···················· | ···················· | ···················· | ···················· | ···················· | ···················· | |
| D | 4048 | hCoV-19/Scotland/EDB6579/2020\|EPI_ISL_478261\|2020- | ···················· | ···················· | ···················· | ···················· | ···················· | ···················· | |
| D | 4049 | hCoV-19/Scotland/EDB6581/2020\|EPI_ISL_478262\|2020- | ···················· | ···················· | ···················· | ···················· | ···················· | ···················· | |
| D | 4050 | hCoV-19/Scotland/EDB6582/2020\|EPI_ISL_478263\|2020- | ···················· | ······A············· | ···················· | ···················· | ···················· | ···················· | |
| D | 4051 | hCoV-19/Scotland/EDB6584/2020\|EPI_ISL_478265\|2020- | ···················· | ···················· | ···················· | ···················· | ···················· | ···················· | |

| | | | | | | | | |
|---|---|---|---|---|---|---|---|---|
| D | 4052 | hCoV-19/Scotland/EDB6585/2020\|EPI_ISL_478266\|2020- | ···················· | ······A············· | ···················· | ···················· | ···················· | ···················· |
| D | 4053 | hCoV-19/Scotland/EDB6586/2020\|EPI_ISL_478267\|2020- | ···················· | ···················· | ···················· | ···················· | ···················· | ···················· |
| D | 4054 | hCoV-19/Scotland/EDB6587/2020\|EPI_ISL_478268\|2020- | ···················· | ···················· | ···················· | ···················· | ···················· | ···················· |
| D | 4055 | hCoV-19/Scotland/EDB6588/2020\|EPI_ISL_478269\|2020- | ···················· | ···················· | ···················· | ···················· | ···················· | ···················· |
| D | 4056 | hCoV-19/Scotland/EDB6590/2020\|EPI_ISL_478270\|2020- | ···················· | ···················· | ···················· | ···················· | ···················· | ···················· |
| D | 4057 | hCoV-19/England/LCST-24D675F/2020\|EPI_ISL_478289\|2 | ···················· | ···················· | ···················· | ···················· | ···················· | ···················· |
| D | 4058 | hCoV-19/England/LCST-24D676E/2020\|EPI_ISL_478290\|2 | ···················· | ···················· | ···················· | ···················· | ···················· | ···················· |
| D | 4059 | hCoV-19/England/LCST-24D67AA/2020\|EPI_ISL_478292\|2 | ···················· | ···················· | ···················· | ···················· | ···················· | ···················· |
| D | 4060 | hCoV-19/England/LCST-24D67B9/2020\|EPI_ISL_478293\|2 | ···················· | ···················· | ···················· | ···················· | ···················· | ···················· |
| D | 4061 | hCoV-19/England/LCST-24D67C8/2020\|EPI_ISL_478294\|2 | ···················· | ···················· | ···················· | ···················· | ···················· | ···················· |
| D | 4062 | hCoV-19/England/LCST-24D67D7/2020\|EPI_ISL_478295\|2 | ···················· | ···················· | ···················· | ···················· | ···················· | ···················· |
| D | 4063 | hCoV-19/England/LCST-24D67E6/2020\|EPI_ISL_478296\|2 | ···················· | ···················· | ···················· | ···················· | ···················· | ···················· |
| D | 4064 | hCoV-19/England/LCST-24D67F5/2020\|EPI_ISL_478297\|2 | ···················· | ···················· | ···················· | ···················· | ···················· | ···················· |
| D | 4065 | hCoV-19/England/LCST-24D6801/2020\|EPI_ISL_478298\|2 | ···················· | ···················· | ···················· | ···················· | ···················· | ···················· |
| D | 4066 | hCoV-19/England/LCST-24D6810/2020\|EPI_ISL_478299\|2 | ···················· | ···················· | ···················· | ···················· | ···················· | ···················· |
| D | 4067 | hCoV-19/India/MH-NIV-45071/2020\|EPI_ISL_479547\|202 | ···················· | ···················· | ···················· | ···················· | ···················· | ···················· |
| D | 4069 | hCoV-19/India/MH-NIV-46317/2020\|EPI_ISL_479550\|202 | ···················· | ···················· | ···················· | ···················· | ···················· | ···················· |
| D | 4070 | hCoV-19/India/MH-NIV-46348/2020\|EPI_ISL_479551\|202 | ···················· | ···················· | ···················· | ···················· | ···················· | ···················· |
| D | 4073 | hCoV-19/India/MH-NIV-64463/2020\|EPI_ISL_479554\|202 | ···················· | ···················· | ···················· | ···················· | ···················· | ···················· |
| D | 4074 | hCoV-19/India/MH-NIV-64478/2020\|EPI_ISL_479555\|202 | ···················· | ···················· | ···················· | ···················· | ···················· | ···················· |
| D | 4075 | hCoV-19/India/MH-NIV-64850/2020\|EPI_ISL_479556\|202 | ···················· | ···················· | ···················· | ···················· | ···················· | ···················· |
| D | 4076 | hCoV-19/India/MH-NIV-64877/2020\|EPI_ISL_479557\|202 | ···················· | ···················· | ···················· | ···················· | ···················· | ···················· |
| D | 4077 | hCoV-19/Singapore/512/2020\|EPI_ISL_479590\|2020-05- | ···················· | ···················· | ···················· | ···················· | ···················· | ···················· |
| D | 4078 | hCoV-19/Singapore/513/2020\|EPI_ISL_479591\|2020-05- | ···················· | ···················· | ···················· | ···················· | ···················· | ···················· |
| D | 4079 | hCoV-19/Singapore/514/2020\|EPI_ISL_479592\|2020-05- | ···················· | ···················· | ···················· | ···················· | ···················· | ···················· |
| D | 4080 | hCoV-19/Singapore/515/2020\|EPI_ISL_479593\|2020-05- | ···················· | ···················· | ···················· | ···················· | ···················· | ···················· |
| D | 4081 | hCoV-19/Singapore/516/2020\|EPI_ISL_479594\|2020-05- | ···················· | ···················· | ···················· | ···················· | ···················· | ···················· |
| D | 4082 | hCoV-19/Singapore/517/2020\|EPI_ISL_479595\|2020-05- | ···················· | ···················· | ···················· | ···················· | ···················· | ···················· |
| D | 4083 | hCoV-19/Singapore/518/2020\|EPI_ISL_479596\|2020-04- | ···················· | ···················· | ···················· | ···················· | ···················· | ···················· |
| D | 4084 | hCoV-19/Singapore/519/2020\|EPI_ISL_479597\|2020-04- | ···················· | ···················· | ···················· | ···················· | ···················· | ···················· |
| D | 4085 | hCoV-19/Singapore/520/2020\|EPI_ISL_479598\|2020-04- | ···················· | ···················· | ···················· | ···················· | ···················· | ···················· |
| D | 4086 | hCoV-19/Singapore/521/2020\|EPI_ISL_479599\|2020-04- | ···················· | ···················· | ···················· | ···················· | ···················· | ···················· |
| D | 4087 | hCoV-19/Bangladesh/DNAS-CPH-isl-479/2020\|EPI_ISL_6 | ···················· | ···················· | ···················· | ···················· | ···················· | ···················· |
| D | 4088 | hCoV-19/Bangladesh/DNAS-CPH-isl-486/2020\|EPI_ISL_6 | ···················· | ···················· | ···················· | ···················· | ···················· | ···················· |
| D | 4089 | hCoV-19/Bangladesh/DNAS-CPH-isl-430/2020\|EPI_ISL_6 | ···················· | ···················· | ···················· | ···················· | ···················· | ···················· |
| D | 4090 | hCoV-19/Bangladesh/DNAS-CPH-isl-510/2020\|EPI_ISL_6 | ···················· | ···················· | ···················· | ···················· | ···················· | ···················· |
| D | 4091 | hCoV-19/Bangladesh/DNAS-CPH-isl-506/2020\|EPI_ISL_6 | ···················· | ···················· | ···················· | ···················· | ···················· | ···················· |
| D | 4092 | hCoV-19/Bangladesh/DNAS-CPH-isl-438/2020\|EPI_ISL_6 | ···················· | ···················· | ···················· | ···················· | ···················· | ···················· |
| D | 4093 | hCoV-19/Bangladesh/DNAS-CPH-isl-525/2020\|EPI_ISL_6 | ···················· | ···················· | ···················· | ···················· | ···················· | ···················· |
| D | 4094 | hCoV-19/Bangladesh/DNAS-CPH-isl-515/2020\|EPI_ISL_6 | ···················· | ···················· | ···················· | ···················· | ···················· | ···················· |
| D | 4095 | hCoV-19/Bangladesh/DNAS-BSH-isl-1/2020\|EPI_ISL_605 | ···················· | ···················· | ···················· | ···················· | ···················· | ···················· |
| D | 4096 | hCoV-19/Bangladesh/DNAS-RAB-isl-88/2020\|EPI_ISL_60 | ···················· | ···················· | ···················· | ···················· | ···················· | ···················· |
| D | 4097 | hCoV-19/USA/MI-MDHHS-SC20624/2020\|EPI_ISL_462706\|2 | ···················· | ···················· | ···················· | ···················· | ···················· | ···················· |
| D | 4098 | hCoV-19/England/20119073404/2020\|EPI_ISL_465100\|20 | ···················· | ···················· | ···················· | ···················· | ···················· | ···················· |
| D | 4100 | hCoV-19/England/20119075504/2020\|EPI_ISL_465104\|20 | ···················· | ···················· | ···················· | ···················· | ·····N·············· | ···················· |
| D | 4101 | hCoV-19/England/20119076104/2020\|EPI_ISL_465105\|20 | ···················· | ···················· | ···················· | ···················· | ···················· | ···················· |
| D | 4102 | hCoV-19/England/20119078604/2020\|EPI_ISL_465108\|20 | ···················· | ···················· | ···················· | ···················· | ···················· | ···················· |
| D | 4103 | hCoV-19/England/20119085304/2020\|EPI_ISL_465115\|20 | ···················· | ···················· | ···················· | ···················· | ···················· | ···················· |
| D | 4104 | hCoV-19/England/20119087104/2020\|EPI_ISL_465120\|20 | ···················· | ···················· | ···················· | ···················· | ···················· | ···················· |
| D | 4105 | hCoV-19/England/20119090204/2020\|EPI_ISL_465125\|20 | ···················· | ···················· | ···················· | ···················· | ···················· | ···················· |
| D | 4109 | hCoV-19/England/20120000104/2020\|EPI_ISL_465136\|20 | ···················· | ···················· | ···················· | ···················· | ···················· | ···················· |
| D | 4110 | hCoV-19/England/20120004504/2020\|EPI_ISL_465143\|20 | ···················· | ···················· | ···················· | ···················· | ···················· | ···················· |
| D | 4111 | hCoV-19/England/20120007504/2020\|EPI_ISL_465145\|20 | ···················· | ···················· | ···················· | ···················· | ··N················· | ···················· |
| D | 4114 | hCoV-19/England/20156002504/2020\|EPI_ISL_466286\|20 | ···················· | ···················· | ···················· | ···················· | ···················· | ···················· |
| D | 4117 | hCoV-19/England/20156004604/2020\|EPI_ISL_466293\|20 | ···················· | ···················· | ···················· | ···················· | ···················· | ···················· |
| D | 4118 | hCoV-19/England/20156033304/2020\|EPI_ISL_466298\|20 | ···················· | ···················· | ···················· | ···················· | ···················· | ···················· |
| D | 4119 | hCoV-19/England/20156033404/2020\|EPI_ISL_466299\|20 | ···················· | ···················· | ···················· | ···················· | ···················· | ···················· |
| D | 4120 | hCoV-19/England/201620057/2020\|EPI_ISL_466400\|2020 | ···················· | ···················· | ···················· | ···················· | ···················· | ···················· |
| D | 4121 | hCoV-19/England/201620058/2020\|EPI_ISL_466401\|2020 | ···················· | ···················· | ···················· | ···················· | ···················· | ···················· |
| D | 4123 | hCoV-19/England/20162010504/2020\|EPI_ISL_466409\|20 | ···················· | ···················· | ···················· | ···················· | ···················· | ···················· |
| D | 4124 | hCoV-19/England/20162011104/2020\|EPI_ISL_466410\|20 | ···················· | ···················· | ···················· | ···················· | ···················· | ···················· |
| D | 4126 | hCoV-19/England/201620657/2020\|EPI_ISL_466432\|2020 | ···················· | ···················· | ···················· | ···················· | ···················· | ···················· |
| D | 4127 | hCoV-19/England/20164005404/2020\|EPI_ISL_466434\|20 | ···················· | ···················· | ···················· | ···················· | ···················· | ···················· |
| D | 4128 | hCoV-19/England/201640075/2020\|EPI_ISL_466436\|2020 | ···················· | ···················· | ···················· | ···················· | ···················· | ···················· |
| D | 4129 | hCoV-19/England/20164013504/2020\|EPI_ISL_466445\|20 | ···················· | ···················· | ···················· | ···················· | ···················· | ···················· |
| D | 4130 | hCoV-19/England/20164016004/2020\|EPI_ISL_466447\|20 | ···················· | ···················· | ···················· | ···················· | ···················· | ···················· |
| D | 4131 | hCoV-19/England/201640252/2020\|EPI_ISL_466452\|2020 | ···················· | ···················· | ···················· | ···················· | ···················· | ···················· |
| D | 4132 | hCoV-19/England/20164056304/2020\|EPI_ISL_466457\|20 | ···················· | ···················· | ···················· | ···················· | ···················· | ···················· |
| D | 4133 | hCoV-19/England/20164065904/2020\|EPI_ISL_466460\|20 | ···················· | ·········R·········· | ···················· | ···················· | ···················· | ···················· |
| D | 4134 | hCoV-19/England/20164066404/2020\|EPI_ISL_466462\|20 | ···················· | ···················· | ···················· | ···················· | ···················· | ···················· |
| D | 4136 | hCoV-19/England/20164072104/2020\|EPI_ISL_466472\|20 | ···T················ | ···················· | ···················· | ···················· | ···················· | ···················· |
| D | 4137 | hCoV-19/England/20164072304/2020\|EPI_ISL_466474\|20 | ···················· | ···················· | ···················· | ···················· | ···················· | ···················· |
| D | 4138 | hCoV-19/England/BRIS-131393/2020\|EPI_ISL_467708\|20 | ···················· | ···················· | ···················· | ···················· | ···················· | ···················· |
| D | 4139 | hCoV-19/England/BRIS-13116C/2020\|EPI_ISL_467713\|20 | ···················· | ···················· | ···················· | ···················· | ···················· | ···················· |
| D | 4140 | hCoV-19/England/BRIS-131375/2020\|EPI_ISL_467718\|20 | ···················· | ···················· | ···················· | ···················· | ···················· | ···················· |
| D | 4141 | hCoV-19/Russia/CRIE163844/2020\|EPI_ISL_467774\|2020 | ···················· | ···················· | ···················· | ···················· | ···················· | ···················· |
| D | 4142 | hCoV-19/Russia/CRIE139642/2020\|EPI_ISL_467775\|2020 | ···················· | ···················· | ···················· | ···················· | ···················· | ···················· |
| D | 4143 | hCoV-19/Bucuresti/ChVir7244/2020\|EPI_ISL_467778\|20 | ···················· | ···················· | ···················· | ···················· | ···················· | ···················· |

| | | | | | | | | |
|---|---|---|---|---|---|---|---|---|
| D | 4144 hCoV-19/Bucuresti/ChVir7245/2020\|EPI_ISL_467779\|20 | .................... | .................... | .................... | .................... | .................... | .................... | |
| D | 4145 hCoV-19/Romania /ChVir7246/2020\|EPI_ISL_467780\|202 | .................... | .................... | .................... | .................... | .................... | .................... | |
| D | 4146 hCoV-19/Romania/ChVir7248/2020\|EPI_ISL_467781\|2020 | .................... | .................... | .................... | .................... | .................... | .................... | |
| D | 4147 hCoV-19/USA/VA-DCLS-0385/2020\|EPI_ISL_467782\|2020- | .................... | .................... | .................... | .................... | .................... | .................... | |
| D | 4148 hCoV-19/USA/VA-DCLS-0386/2020\|EPI_ISL_467783\|2020- | .................... | .................... | .................... | .................... | .................... | .................... | |
| D | 4149 hCoV-19/USA/VA-DCLS-0387/2020\|EPI_ISL_467784\|2020- | .................... | .................... | .................... | .................... | .................... | .................... | |
| D | 4150 hCoV-19/USA/VA-DCLS-0388/2020\|EPI_ISL_467785\|2020- | .................... | .................... | .................... | .................... | .................... | .................... | |
| D | 4151 hCoV-19/USA/VA-DCLS-0389/2020\|EPI_ISL_467786\|2020- | .................... | .................... | .................... | .................... | .................... | .................... | |
| D | 4152 hCoV-19/USA/VA-DCLS-0390/2020\|EPI_ISL_467787\|2020- | .................... | .................... | .................... | .................... | .................... | .................... | |
| D | 4153 hCoV-19/USA/VA-DCLS-0392/2020\|EPI_ISL_467788\|2020- | .................... | .................... | .................... | .................... | .................... | .................... | |
| D | 4154 hCoV-19/USA/VA-DCLS-0393/2020\|EPI_ISL_467789\|2020- | .................... | .................... | .................... | .................... | .................... | .................... | |
| D | 4155 hCoV-19/USA/VA-DCLS-0395/2020\|EPI_ISL_467790\|2020- | .................... | .................... | .................... | .................... | .................... | .................... | |
| D | 4156 hCoV-19/USA/VA-DCLS-0396/2020\|EPI_ISL_467791\|2020- | .................... | .................... | .................... | .................... | .................... | .................... | |
| D | 4157 hCoV-19/USA/VA-DCLS-0397/2020\|EPI_ISL_467792\|2020- | .................... | .................... | .................... | .................... | .................... | .................... | |
| D | 4158 hCoV-19/USA/VA-DCLS-0398/2020\|EPI_ISL_467793\|2020- | .................... | .................... | .................... | .................... | .................... | .........A.......... | |
| D | 4159 hCoV-19/USA/VA-DCLS-0399/2020\|EPI_ISL_467794\|2020- | .................... | .................... | .................... | .................... | .................... | .................... | |
| D | 4160 hCoV-19/USA/VA-DCLS-0400/2020\|EPI_ISL_467795\|2020- | .................... | .................... | .................... | .................... | .................... | .................... | |
| D | 4161 hCoV-19/USA/VA-DCLS-0401/2020\|EPI_ISL_467796\|2020- | .................... | .................... | .................... | .................... | .................... | .................... | |
| D | 4162 hCoV-19/USA/VA-DCLS-0402/2020\|EPI_ISL_467797\|2020- | .................... | .................... | .................... | .................... | .................... | .................... | |
| D | 4163 hCoV-19/USA/VA-DCLS-0403/2020\|EPI_ISL_467798\|2020- | .................... | .................... | .................... | .................... | .................... | .................... | |
| D | 4164 hCoV-19/USA/VA-DCLS-0404/2020\|EPI_ISL_467799\|2020- | .................... | .................... | .................... | .................... | .................... | .................... | |
| D | 4165 hCoV-19/England/NORT-283BB7/2020\|EPI_ISL_472249\|20 | .................... | .................... | .................... | .................... | .................... | .................... | |
| D | 4166 hCoV-19/England/NORT-283DEE/2020\|EPI_ISL_472255\|20 | .................... | .................... | .................... | .................... | .................... | .................... | |
| D | 4167 hCoV-19/England/NORT-283E27/2020\|EPI_ISL_472256\|20 | .................... | .................... | .................... | .................... | .................... | .................... | |
| D | 4169 hCoV-19/England/NORW-EB074/2020\|EPI_ISL_472296\|202 | .................... | .................... | .................... | .................... | N.N................. | .................... | |
| D | 4170 hCoV-19/Scotland/CVR2599/2020\|EPI_ISL_473524\|2020- | .................... | .................... | .................... | .................... | .................... | .................... | |
| D | 4171 hCoV-19/Scotland/CVR2782/2020\|EPI_ISL_473563\|2020- | .................... | .................... | .................... | .................... | .................... | .................... | |
| D | 4173 hCoV-19/Scotland/CVR2787/2020\|EPI_ISL_473566\|2020- | .................... | .................... | .................... | .................... | .................... | .................... | |
| D | 4174 hCoV-19/Scotland/CVR2788/2020\|EPI_ISL_473567\|2020- | .................... | ...................A | .................... | .................... | .................... | .................... | 1 |
| D | 4175 hCoV-19/Scotland/CVR2790/2020\|EPI_ISL_473568\|2020- | .................... | .................... | .................... | .................... | .................... | .................... | |
| D | 4176 hCoV-19/Scotland/CVR2791/2020\|EPI_ISL_473569\|2020- | .................... | ...................A | .................... | .................... | .................... | .................... | 1 |
| D | 4177 hCoV-19/Scotland/CVR2792/2020\|EPI_ISL_473570\|2020- | .................... | .................... | .................... | .................... | .................... | .................... | |
| D | 4178 hCoV-19/Scotland/CVR2804/2020\|EPI_ISL_473572\|2020- | .................... | ...................A | .................... | .................... | .................... | .................... | 1 |
| D | 4179 hCoV-19/Scotland/CVR2806/2020\|EPI_ISL_473573\|2020- | .................... | .................... | .................... | .................... | .................... | .................... | |
| D | 4180 hCoV-19/Scotland/CVR2807/2020\|EPI_ISL_473574\|2020- | .................... | ...................A | .................... | .................... | .................... | .................... | 1 |
| D | 4181 hCoV-19/Scotland/CVR2808/2020\|EPI_ISL_473575\|2020- | .................... | .................... | .................... | .................... | .................... | .................... | |
| D | 4182 hCoV-19/Scotland/CVR2809/2020\|EPI_ISL_473576\|2020- | .................... | .................... | .................... | .................... | .................... | .................... | |
| D | 4183 hCoV-19/Scotland/CVR2811/2020\|EPI_ISL_473577\|2020- | .................... | .................... | .................... | .................... | .................... | .................... | |
| D | 4184 hCoV-19/Scotland/CVR2813/2020\|EPI_ISL_473578\|2020- | .................... | .................... | .................... | .................... | .................... | .................... | |
| D | 4185 hCoV-19/Scotland/CVR2815/2020\|EPI_ISL_473579\|2020- | .................... | .................... | .................... | .................... | .................... | .................... | |
| D | 4186 hCoV-19/Scotland/CVR2816/2020\|EPI_ISL_473580\|2020- | .................... | .................... | .................... | .................... | .................... | .................... | |
| D | 4187 hCoV-19/Scotland/CVR2818/2020\|EPI_ISL_473581\|2020- | .................... | .................... | .................... | .................... | .................... | .................... | |
| D | 4188 hCoV-19/Scotland/CVR2820/2020\|EPI_ISL_473582\|2020- | .................... | ..............C..... | .................... | .................... | .................... | .................... | |
| D | 4189 hCoV-19/Scotland/CVR2821/2020\|EPI_ISL_473583\|2020- | .................... | .................... | .................... | .................... | .................... | .................... | |
| D | 4190 hCoV-19/Scotland/CVR2822/2020\|EPI_ISL_473584\|2020- | .................... | .................... | .................... | .................... | .................... | .................... | |
| D | 4191 hCoV-19/Scotland/CVR2823/2020\|EPI_ISL_473585\|2020- | .................... | .A.................. | .................... | .................... | .................... | .................... | |
| D | 4192 hCoV-19/Scotland/CVR2824/2020\|EPI_ISL_473586\|2020- | .................... | .................... | .................... | .................... | .................... | .................... | |
| D | 4193 hCoV-19/Scotland/CVR2825/2020\|EPI_ISL_473587\|2020- | .................... | .................... | .................... | .................... | .................... | .................... | |
| D | 4194 hCoV-19/Scotland/CVR2826/2020\|EPI_ISL_473588\|2020- | .................... | ...................A | .................... | .................... | .................... | .................... | 1 |
| D | 4195 hCoV-19/Scotland/CVR2827/2020\|EPI_ISL_473589\|2020- | .................... | .................... | .................... | .................... | .................... | .................... | |
| D | 4196 hCoV-19/Scotland/CVR2828/2020\|EPI_ISL_473590\|2020- | .................... | .A.................. | .................... | .................... | .................... | .................... | |
| D | 4198 hCoV-19/Scotland/CVR2847/2020\|EPI_ISL_473592\|2020- | .................... | ..........A......... | .................... | .................... | .................... | .................... | |
| D | 4199 hCoV-19/Scotland/CVR2848/2020\|EPI_ISL_473593\|2020- | .................... | ...................A | .................... | .................... | .................... | .................... | 1 |
| D | 4201 hCoV-19/Scotland/CVR2850/2020\|EPI_ISL_473595\|2020- | .................... | .................... | .................... | .................... | .................... | .................... | |
| D | 4202 hCoV-19/Scotland/CVR2851/2020\|EPI_ISL_473596\|2020- | .................... | .................... | .................... | .................... | .................... | .................... | |
| D | 4203 hCoV-19/Scotland/CVR2854/2020\|EPI_ISL_473597\|2020- | .................... | .................... | .................... | .................... | .................... | .................... | |
| D | 4204 hCoV-19/Scotland/CVR2855/2020\|EPI_ISL_473598\|2020- | .................... | ..............C..... | .................... | .................... | .................... | .................... | |
| D | 4205 hCoV-19/Scotland/CVR2856/2020\|EPI_ISL_473599\|2020- | .................... | .................... | .................... | .................... | .................... | .................... | |
| D | 4210 hCoV-19/Spain/AN-IBV-002887/2020\|EPI_ISL_474897\|20 | .................... | .................... | .................... | .................... | .................... | .................... | |
| D | 4212 hCoV-19/Canada/BC_07917795/2020\|EPI_ISL_477100\|202 | .................... | .................... | .................... | .................... | .................... | .................... | |
| D | 4213 hCoV-19/Canada/BC_07920084/2020\|EPI_ISL_477101\|202 | .................... | .................... | .................... | .................... | .................... | .................... | |
| D | 4214 hCoV-19/Canada/BC_07924910/2020\|EPI_ISL_477102\|202 | .................... | .................... | .................... | .................... | .................... | .................... | |
| D | 4215 hCoV-19/Canada/BC_07925997/2020\|EPI_ISL_477103\|202 | .................... | .................... | .................... | .................... | .................... | .................... | |
| D | 4216 hCoV-19/Canada/BC_09009433/2020\|EPI_ISL_477105\|202 | .................... | .................... | .................... | .................... | .................... | .................... | |
| D | 4217 hCoV-19/Canada/BC_09014362/2020\|EPI_ISL_477106\|202 | .................... | .................... | .................... | .................... | .................... | .................... | |
| D | 4218 hCoV-19/Canada/BC_09021025/2020\|EPI_ISL_477107\|202 | .................... | .................... | .................... | .................... | .................... | .................... | |
| D | 4219 hCoV-19/Canada/BC_09023420/2020\|EPI_ISL_477108\|202 | .................... | .................... | .................... | .................... | .................... | .................... | |
| D | 4220 hCoV-19/Canada/BC_09025513/2020\|EPI_ISL_477109\|202 | .................... | .................... | .................... | .................... | .................... | .................... | |
| D | 4221 hCoV-19/Canada/BC_09026743/2020\|EPI_ISL_477110\|202 | .................... | .................... | .................... | .................... | .................... | .................... | |
| D | 4222 hCoV-19/Canada/BC_09029034/2020\|EPI_ISL_477111\|202 | .................... | .................... | .................... | .................... | .................... | .................... | |
| D | 4223 hCoV-19/Canada/BC_09035190/2020\|EPI_ISL_477112\|202 | .................... | .................... | .................... | .................... | .................... | .................... | |
| D | 4224 hCoV-19/Canada/BC_09040534/2020\|EPI_ISL_477113\|202 | .................... | .................... | .................... | .................... | .................... | .................... | |
| D | 4225 hCoV-19/Canada/BC_10600259/2020\|EPI_ISL_477114\|202 | .................... | .................... | .................... | .................... | .................... | .................... | |
| D | 4226 hCoV-19/Canada/BC_10600634/2020\|EPI_ISL_477115\|202 | .................... | .................... | .................... | .................... | .................... | .................... | |
| D | 4227 hCoV-19/Canada/BC_10601737/2020\|EPI_ISL_477117\|202 | .................... | .................... | .................... | .................... | .................... | .................... | |
| D | 4228 hCoV-19/Canada/BC_10603886/2020\|EPI_ISL_477118\|202 | .................... | .................... | .................... | .................... | .................... | .................... | |
| D | 4229 hCoV-19/Canada/BC_10616731/2020\|EPI_ISL_477119\|202 | .................... | .................... | .................... | .................... | .................... | .................... | |
| D | 4230 hCoV-19/Bangladesh/CHRF-0012/2020\|EPI_ISL_477125\|2 | .................... | .................... | .................... | .................... | .................... | .................... | |

| | | | | | | | | |
|---|---|---|---|---|---|---|---|---|
| D | 4231 hCoV-19/Bangladesh/CHRF-0013/2020\|EPI_ISL_477126\|2 | ···················· | ···················· | ···················· | ···················· | ···················· | ···················· | |
| D | 4232 hCoV-19/Scotland/EDB6099/2020\|EPI_ISL_478221\|2020- | ···················· | ···················A | ···················· | ···················· | ···················· | ···················· | 1 |
| D | 4233 hCoV-19/Scotland/EDB6109/2020\|EPI_ISL_478227\|2020- | ···················· | ···················A | ···················· | ···················· | ···················· | ···················· | 1 |
| D | 4234 hCoV-19/Scotland/EDB6118/2020\|EPI_ISL_478230\|2020- | ···················· | ···················A | ···················· | ···················· | ···················· | ···················· | 1 |
| D | 4235 hCoV-19/Scotland/EDB6121/2020\|EPI_ISL_478232\|2020- | ···················· | ···················A | ···················· | ···················· | ···················· | ···················· | 1 |
| D | 4236 hCoV-19/Scotland/EDB6123/2020\|EPI_ISL_478233\|2020- | ···················· | ···················A | ···················· | ···················· | ···················· | ···················· | 1 |
| D | 4237 hCoV-19/Scotland/EDB6515/2020\|EPI_ISL_478249\|2020- | ···················· | ···················· | ···················· | ···················· | ···················· | ···················· | |
| D | 4238 hCoV-19/Scotland/EDB6517/2020\|EPI_ISL_478250\|2020- | ···················· | ···················· | ···················· | ···················· | ···················· | ···················· | |
| D | 4239 hCoV-19/Scotland/EDB6530/2020\|EPI_ISL_478251\|2020- | ···················· | ···················· | ···················· | ···················· | ···················· | ···················· | |
| D | 4240 hCoV-19/Scotland/EDB6535/2020\|EPI_ISL_478252\|2020- | ···················· | ···················· | ···················· | ···················· | ···················· | ···················· | |
| D | 4241 hCoV-19/Scotland/EDB6568/2020\|EPI_ISL_478254\|2020- | ···················· | ·····A·············· | ···················· | ···················· | ···················· | ···················· | |
| D | 4242 hCoV-19/Scotland/EDB6569/2020\|EPI_ISL_478255\|2020- | ···················· | ·····A·············· | ···················· | ···················· | ···················· | ···················· | |
| D | 4243 hCoV-19/Scotland/EDB6573/2020\|EPI_ISL_478256\|2020- | ···················· | ···················· | ···················· | ···················· | ···················· | ···················· | |
| D | 4244 hCoV-19/Scotland/EDB6575/2020\|EPI_ISL_478257\|2020- | ···················· | ···················· | ···················· | ···················· | ···················· | ···················· | |
| D | 4245 hCoV-19/Scotland/EDB6576/2020\|EPI_ISL_478258\|2020- | ···················· | ···················· | ···················· | ···················· | ···················· | ···················· | |
| D | 4246 hCoV-19/Scotland/EDB6577/2020\|EPI_ISL_478259\|2020- | ···················· | ···················· | ···················· | ···················· | ···················· | ···················· | |
| D | 4247 hCoV-19/England/LOND-D4278/2020\|EPI_ISL_478404\|202 | ···················· | ···················· | ···················· | ···················· | ···················· | ···················· | |
| D | 4248 hCoV-19/England/LOND-D47C4/2020\|EPI_ISL_478405\|202 | ···················· | ···················· | ···················· | ···················· | ···················· | ···················· | |
| D | 4249 hCoV-19/England/LOND-D4E83/2020\|EPI_ISL_478406\|202 | ···················· | ···················· | ···················· | ···················· | ···················· | ···················· | |
| D | 4250 hCoV-19/England/LOND-D5DD0/2020\|EPI_ISL_478410\|202 | ···················· | ···················· | ···················· | ···················· | ···················· | ···················· | |
| D | 4251 hCoV-19/England/LOND-D6BA8/2020\|EPI_ISL_478411\|202 | ···················· | ···················· | ···················· | ···················· | ···················· | ···················· | |
| D | 4252 hCoV-19/England/LOND-D6BB7/2020\|EPI_ISL_478412\|202 | ···················· | ···················· | ···················· | ···················· | ···················· | ···················· | |
| D | 4253 hCoV-19/England/LOND-D6BC6/2020\|EPI_ISL_478413\|202 | ···················· | ···················· | ···················· | ···················· | ···················· | ···················· | |
| D | 4254 hCoV-19/England/LOND-D6BD5/2020\|EPI_ISL_478414\|202 | ···················· | ···················· | ···················· | ···················· | ···················· | ···················· | |
| D | 4255 hCoV-19/England/LOND-D6C3C/2020\|EPI_ISL_478415\|202 | ···················· | ···················· | ···················· | ···················· | ···················· | ···················· | |
| D | 4256 hCoV-19/England/LOND-D6C87/2020\|EPI_ISL_478417\|202 | ···················· | ···················· | ···················· | ···················· | ···················· | ···················· | |
| D | 4257 hCoV-19/England/LOND-D6C96/2020\|EPI_ISL_478418\|202 | ···················· | ···················· | ···················· | ···················· | ···················· | ···················· | |
| D | 4258 hCoV-19/England/LOND-D6CA5/2020\|EPI_ISL_478419\|202 | ···················· | ···················· | ···················· | ···················· | ···················· | ···················· | |
| D | 4259 hCoV-19/England/LOND-D6CB4/2020\|EPI_ISL_478420\|202 | ···················· | ···················· | ···················· | ···················· | ···················· | ···················· | |
| D | 4260 hCoV-19/England/LOND-D6CC3/2020\|EPI_ISL_478421\|202 | ···················· | ···················· | ···················· | ···················· | ···················· | ···················· | |
| D | 4261 hCoV-19/England/LOND-D6CD2/2020\|EPI_ISL_478422\|202 | ···················· | ···················· | ···················· | ···················· | ···················· | ···················· | |
| D | 4262 hCoV-19/England/LOND-D6D0C/2020\|EPI_ISL_478424\|202 | ···················· | ···················· | ···················· | ···················· | ···················· | ···················· | |
| D | 4263 hCoV-19/England/LOND-D6D2A/2020\|EPI_ISL_478426\|202 | ···················· | ···················· | ···················· | ···················· | ···················· | ···················· | |
| D | 4264 hCoV-19/England/LOND-D6D39/2020\|EPI_ISL_478427\|202 | ···················· | ···················· | ···················· | ···················· | ···················· | ···················· | |
| D | 4265 hCoV-19/England/LOND-D6D48/2020\|EPI_ISL_478428\|202 | ···················· | ···················· | ···················· | ···················· | ···················· | ···················· | |
| D | 4266 hCoV-19/England/LOND-D6D66/2020\|EPI_ISL_478429\|202 | ···················· | ···················· | ···················· | ···················· | ···················· | ···················· | |
| D | 4267 hCoV-19/England/LOND-D6D75/2020\|EPI_ISL_478430\|202 | ···················· | ···················· | ···················· | ···················· | ···················· | ···················· | |
| D | 4268 hCoV-19/England/LOND-D6DB1/2020\|EPI_ISL_478433\|202 | ···················· | ···················· | ···················· | ···················· | ···················· | ···················· | |
| D | 4269 hCoV-19/England/LOND-D6DC0/2020\|EPI_ISL_478434\|202 | ···················· | ···················· | ···················· | ···················· | ···················· | ···················· | |
| D | 4270 hCoV-19/England/LOND-D6DDF/2020\|EPI_ISL_478435\|202 | ···················· | ···················· | ···················· | ···················· | ···················· | ···················· | |
| D | 4271 hCoV-19/England/LOND-D6E45/2020\|EPI_ISL_478438\|202 | ···················· | ···················· | ···················· | ···················· | ···················· | ···················· | |
| D | 4272 hCoV-19/England/LOND-D6E63/2020\|EPI_ISL_478440\|202 | ···················· | ···················· | ···················· | ···················· | ···················· | ···················· | |
| D | 4273 hCoV-19/England/LOND-D6E72/2020\|EPI_ISL_478441\|202 | ···················· | ···················· | ···················· | ···················· | ···················· | ···················· | |
| D | 4274 hCoV-19/England/LOND-D6EBE/2020\|EPI_ISL_478443\|202 | ···················· | ···················· | ···················· | ···················· | ···················· | ···················· | |
| D | 4275 hCoV-19/England/LOND-D6F42/2020\|EPI_ISL_478444\|202 | ···················· | ···················· | ···················· | ···················· | ···················· | ···················· | |
| D | 4276 hCoV-19/England/LOND-D6F60/2020\|EPI_ISL_478445\|202 | ···················· | ···················· | ···················· | ···················· | ···················· | ···················· | |
| D | 4277 hCoV-19/England/LOND-D6FD9/2020\|EPI_ISL_478450\|202 | ···················· | ···················· | ···················· | ···················· | ···················· | ···················· | |
| D | 4278 hCoV-19/England/LOND-D6FF7/2020\|EPI_ISL_478451\|202 | ···················· | ···················· | ···················· | ···················· | ···················· | ···················· | |
| D | 4279 hCoV-19/England/LOND-D7002/2020\|EPI_ISL_478452\|202 | ···················· | ···················· | ···················· | ···················· | ···················· | ···················· | |
| D | 4280 hCoV-19/England/LOND-D7011/2020\|EPI_ISL_478453\|202 | ···················· | ···················· | ···················· | ···················· | ···················· | ···················· | |
| D | 4281 hCoV-19/England/LOND-D703F/2020\|EPI_ISL_478454\|202 | ···················· | ···················· | ···················· | ···················· | ···················· | ···················· | |
| D | 4282 hCoV-19/England/LOND-D704E/2020\|EPI_ISL_478455\|202 | ···················· | ···················· | ···················· | ···················· | ···················· | ···················· | |
| D | 4283 hCoV-19/England/LOND-D705D/2020\|EPI_ISL_478456\|202 | ···················· | ···················· | ···················· | ···················· | ···················· | ···················· | |
| D | 4284 hCoV-19/England/LOND-D706C/2020\|EPI_ISL_478457\|202 | ···················· | ···················· | ···················· | ···················· | ···················· | ···················· | |
| D | 4286 hCoV-19/India/MH-NIV-64905/2020\|EPI_ISL_479559\|202 | ···················· | ···················· | ···················· | ···················· | ···················· | ···················· | |
| D | 4287 hCoV-19/India/MH-NIV-64908/2020\|EPI_ISL_479560\|202 | ···················· | ···················· | ···················· | ···················· | ···················· | ···················· | |
| D | 4288 hCoV-19/India/MH-NIV-65282/2020\|EPI_ISL_479562\|202 | ···················· | ···················· | ···················· | ···················· | ···················· | ···················· | |
| D | 4289 hCoV-19/India/MH-NIV-65417/2020\|EPI_ISL_479563\|202 | ···················· | ···················· | ···················· | ···················· | ···················· | ···················· | |
| D | 4290 hCoV-19/India/MH-NIV-65433/2020\|EPI_ISL_479564\|202 | ···················· | ···················· | ···················· | ···················· | ···················· | ···················· | |
| D | 4291 hCoV-19/India/MH-NIV-65813/2020\|EPI_ISL_479565\|202 | ···················· | ···················· | ···················· | ···················· | ···················· | ···················· | |
| D | 4292 hCoV-19/India/MH-NIV-65829/2020\|EPI_ISL_479566\|202 | ···················· | ···················· | ···················· | ···················· | ···················· | ···················· | |
| D | 4293 hCoV-19/India/MH-NIV-65835/2020\|EPI_ISL_479567\|202 | ···················· | ···················· | ···················· | ···················· | ···················· | ···················· | |
| D | 4294 hCoV-19/India/MH-NIV-65847/2020\|EPI_ISL_479568\|202 | ···················· | ···················· | ···················· | ···················· | ···················· | ···················· | |
| D | 4295 hCoV-19/India/MH-NIV-65877/2020\|EPI_ISL_479570\|202 | ···················· | ···················· | ···················· | ···················· | ···················· | ···················· | |
| D | 4296 hCoV-19/India/MH-NIV-65880/2020\|EPI_ISL_479571\|202 | ···················· | ···················· | ···················· | ···················· | ···················· | ···················· | |
| D | 4297 hCoV-19/India/MH-NIV-SARI-4375/2020\|EPI_ISL_479572 | ···················· | ···················· | ···················· | ···················· | ···················· | ···················· | |
| D | 4298 hCoV-19/Singapore/496/2020\|EPI_ISL_479574\|2020-04- | ···················· | ···················· | ···················· | ···················· | ···················· | ···················· | |
| D | 4299 hCoV-19/Singapore/497/2020\|EPI_ISL_479575\|2020-04- | ···················· | ···················· | ···················· | ···················· | ···················· | ···················· | |
| D | 4300 hCoV-19/Singapore/498/2020\|EPI_ISL_479576\|2020-04- | ···················· | ···················· | ···················· | ···················· | ···················· | ···················· | |
| D | 4301 hCoV-19/Singapore/499/2020\|EPI_ISL_479577\|2020-04- | ···················· | ···················· | ···················· | ···················· | ···················· | ···················· | |
| D | 4302 hCoV-19/Singapore/500/2020\|EPI_ISL_479578\|2020-04- | ···················· | ···················· | ···················· | ···················· | ···················· | ···················· | |
| D | 4303 hCoV-19/Singapore/502/2020\|EPI_ISL_479580\|2020-04- | ···················· | ···················· | ···················· | ···················· | ···················· | ···················· | |
| D | 4304 hCoV-19/Singapore/503/2020\|EPI_ISL_479581\|2020-04- | ···················· | ···················· | ···················· | ···················· | ···················· | ···················· | |
| D | 4305 hCoV-19/Singapore/504/2020\|EPI_ISL_479582\|2020-04- | ···················· | ···················· | ···················· | ···················· | ···················· | ···················· | |
| D | 4306 hCoV-19/Singapore/505/2020\|EPI_ISL_479583\|2020-06- | ···················· | ···················· | ···················· | ···················· | ···················· | ···················· | |
| D | 4307 hCoV-19/Singapore/506/2020\|EPI_ISL_479584\|2020-05- | ···················· | ···················· | ···················· | ···················· | ···················· | ···················· | |
| D | 4308 hCoV-19/Singapore/507/2020\|EPI_ISL_479585\|2020-05- | ···················· | ···················· | ···················· | ···················· | ···················· | ···················· | |
| D | 4309 hCoV-19/Singapore/508/2020\|EPI_ISL_479586\|2020-05- | ···················· | ···················· | ···················· | ···················· | ···················· | ···················· | |

| | | | | | | | |
|---|---|---|---|---|---|---|---|
| D | 4310 hCoV-19/Singapore/509/2020\|EPI_ISL_479587\|2020-05- | ···················· | ···················· | ····················· | ···················· | ···················· | ···················· |
| D | 4311 hCoV-19/Singapore/510/2020\|EPI_ISL_479588\|2020-05- | ···················· | ···················· | ····················· | ···················· | ···················· | ···················· |
| D | 4312 hCoV-19/Singapore/511/2020\|EPI_ISL_479589\|2020-05- | ···················· | ···················· | ····················· | ···················· | ···················· | ···················· |
| D | 4313 hCoV-19/Egypt/CUNCI-HGC4I004/2020\|EPI_ISL_479700\|2 | ···················· | ···················· | ····················· | ···················· | ···················· | ···················· |
| D | 4314 hCoV-19/Egypt/CUNCI-HGC4I029/2020\|EPI_ISL_479701\|2 | ···················· | ···················· | ····················· | ···················· | ···················· | ···················· |
| D | 4315 hCoV-19/Egypt/CUNCI-HGC4I030/2020\|EPI_ISL_479703\|2 | ···················· | ···················· | ····················· | ···················· | ···················· | ···················· |
| D | 4316 hCoV-19/Egypt/CUNCI-HGC4I033/2020\|EPI_ISL_479704\|2 | ···················· | ··············Y····· | ····················· | ···················· | ···················· | ···················· |
| D | 4317 hCoV-19/Egypt/CUNCI-HGC4I022/2020\|EPI_ISL_479705\|2 | ···················· | ···················· | ····················· | ···················· | ···················· | ···················· |
| D | 4318 hCoV-19/Egypt/CUNCI-HGC4I025/2020\|EPI_ISL_479706\|2 | ···················· | ···················· | ····················· | ···················· | ···················· | ···················· |
| D | 4319 hCoV-19/Egypt/CUNCI-HGC4I031/2020\|EPI_ISL_479707\|2 | ···················· | ···················· | ····················· | ···················· | ···················· | ···················· |
| D | 4320 hCoV-19/Egypt/CUNCI-HGC4I034/2020\|EPI_ISL_479708\|2 | ···················· | ···················· | ····················· | ···················· | ···················· | ···················· |
| D | 4321 hCoV-19/Egypt/CUNCI-HGC5I001/2020\|EPI_ISL_479710\|2 | ···················· | ···················· | ····················· | ···················· | ···················· | ···················· |
| D | 4322 hCoV-19/Egypt/CUNCI-HGC5I003/2020\|EPI_ISL_479711\|2 | ···················· | ···················· | ····················· | ···················· | ···················· | ···················· |
| D | 4323 hCoV-19/Egypt/CUNCI-HGC5I011/2020\|EPI_ISL_479712\|2 | ···················· | ···················· | ····················· | ···················· | ···················· | ···················· |
| D | 4324 hCoV-19/Egypt/CUNCI-HGC5I012/2020\|EPI_ISL_479713\|2 | ···················· | ···················· | ····················· | ···················· | ···················· | ···················· |
| D | 4325 hCoV-19/Egypt/CUNCI-HGC5I014/2020\|EPI_ISL_479714\|2 | ···················· | ···················· | ····················· | ···················· | ···················· | ···················· |
| D | 4326 hCoV-19/Egypt/CUNCI-HGC5I022/2020\|EPI_ISL_479715\|2 | ···················· | ···················· | ····················· | ···················· | ···················· | ···················· |
| D | 4327 hCoV-19/Egypt/CUNCI-HGC5I025/2020\|EPI_ISL_479716\|2 | ···················· | ·R·················· | ····················· | ···················· | ···················· | ···R················ |
| D | 4328 hCoV-19/Egypt/CUNCI-HGC5I033/2020\|EPI_ISL_479717\|2 | ···················· | ···················· | ····················· | ···················· | ···················· | ···················· |
| D | 4329 hCoV-19/Egypt/CUNCI-HGC6I003/2020\|EPI_ISL_479718\|2 | ···················· | ···················· | ····················· | ···················· | ···················· | ···················· |
| D | 4330 hCoV-19/Egypt/CUNCI-HGC6I002/2020\|EPI_ISL_479719\|2 | ···················· | ·········A·········· | ····················· | ···················· | ···················· | ···················· |
| D | 4331 hCoV-19/Egypt/CUNCI-HGC5I034/2020\|EPI_ISL_479720\|2 | ···················· | ···················· | ····················· | ···················· | ···················· | ···················· |
| D | 4332 hCoV-19/Egypt/CUNCI-HGC5I030/2020\|EPI_ISL_479721\|2 | ···················· | ···················· | ····················· | ···················· | ···················· | ···················· |
| D | 4333 hCoV-19/Egypt/CUNCI-HGC6I007/2020\|EPI_ISL_479722\|2 | ···················· | ···················· | ····················· | ···················· | ···················· | ···················· |
| D | 4334 hCoV-19/Egypt/CUNCI-HGC6I033/2020\|EPI_ISL_479723\|2 | ···················· | ·········A·········· | ····················· | ···················· | ···················· | ···················· |
| D | 4335 hCoV-19/Egypt/CUNCI-HGC6I026/2020\|EPI_ISL_479724\|2 | ···················· | ·········A·········· | ····················· | ···················· | ···················· | ···················· |
| D | 4336 hCoV-19/Egypt/CUNCI-HGC6I015/2020\|EPI_ISL_479725\|2 | ···················· | ···················· | ····················· | ···················· | ···················· | ···················· |
| D | 4337 hCoV-19/Egypt/CUNCI-HGC6I009/2020\|EPI_ISL_479726\|2 | ···················· | ·········A·········· | ····················· | ···················· | ···················· | ···················· |
| D | 4338 hCoV-19/Egypt/CUNCI-HGC6I011/2020\|EPI_ISL_479727\|2 | ···················· | ···················· | ····················· | ···················· | ···················· | ···················· |
| D | 4339 hCoV-19/Egypt/CUNCI-HGC4I015/2020\|EPI_ISL_479728\|2 | ···················· | ···················· | ····················· | ···················· | ···················· | ···················· |
| D | 4340 hCoV-19/Egypt/CUNCI-HGC4I023/2020\|EPI_ISL_479729\|2 | ···················· | ···················· | ····················· | ···················· | ···················· | ···················· |
| D | 4341 hCoV-19/Egypt/CUNCI-HGC5I010/2020\|EPI_ISL_479730\|2 | ···················· | ···················· | ····················· | ···················· | ···················· | ···················· |
| D | 4342 hCoV-19/Egypt/CUNCI-HGC5I016/2020\|EPI_ISL_479731\|2 | ···················· | ···················· | ····················· | ···················· | ···················· | ···················· |
| D | 4343 hCoV-19/Egypt/CUNCI-HGC6I032/2020\|EPI_ISL_479733\|2 | ···················· | ·········A·········· | ····················· | ···················· | ···················· | ···················· |
| D | 4344 hCoV-19/Egypt/CUNCI-HGC5I029/2020\|EPI_ISL_479734\|2 | ···················· | ······R············· | ····················· | ···················· | ···················· | ···················· |
| D | 4345 hCoV-19/Egypt/CUNCI-HGC5I026/2020\|EPI_ISL_479735\|2 | ···················· | ···················· | ····················· | ···················· | ···················· | ···················· |
| D | 4346 hCoV-19/Japan/TK-YE611820/2020\|EPI_ISL_486890\|2020 | ···················· | ···················· | ····················· | ···················· | ···················· | ···················· |
| D | 4347 hCoV-19/Japan/TK-YE611821/2020\|EPI_ISL_486891\|2020 | ···················· | ···················· | ····················· | ···················· | ···················· | ···················· |
| D | 4348 hCoV-19/Japan/TK-YV12527/2020\|EPI_ISL_486892\|2020- | ···················· | ···················· | ····················· | ···················· | ···················· | ···················· |
| D | 4349 hCoV-19/Japan/TK-YE612251/2020\|EPI_ISL_486893\|2020 | ···················· | ···················· | ····················· | ···················· | ···················· | ···················· |
| D | 4350 hCoV-19/Japan/TK-YV12689/2020\|EPI_ISL_486894\|2020- | ···················· | ···················· | ····················· | ···················· | ···················· | ···················· |
| D | 4351 hCoV-19/Japan/TK-YE613035/2020\|EPI_ISL_486895\|2020 | ···················· | ···················· | ····················· | ···················· | ···················· | ···················· |
| D | 4352 hCoV-19/Japan/TK-YE613175/2020\|EPI_ISL_486896\|2020 | ···················· | ···················· | ····················· | ···················· | ···················· | ···················· |
| D | 4353 hCoV-19/Canada/BC_22430814/2020\|EPI_ISL_462830\|202 | ···················· | ···················· | ····················· | ···················· | ···················· | ···················· |
| D | 4354 hCoV-19/Canada/BC_22451513/2020\|EPI_ISL_462831\|202 | ···················· | ···················· | ····················· | ···················· | ···················· | ···················· |
| D | 4355 hCoV-19/Canada/BC_22470913/2020\|EPI_ISL_462833\|202 | ···················· | ···················· | ····················· | ···················· | ···················· | ···················· |
| D | 4356 hCoV-19/Canada/BC_22473210/2020\|EPI_ISL_462834\|202 | ···················· | ···················· | ····················· | ···················· | ···················· | ···················· |
| D | 4357 hCoV-19/Canada/BC_22475116/2020\|EPI_ISL_462835\|202 | ···················· | ···················· | ····················· | ···················· | ···················· | ···················· |
| D | 4358 hCoV-19/Canada/BC_22511314/2020\|EPI_ISL_462836\|202 | ···················· | ···················· | ····················· | ···················· | ···················· | ···················· |
| D | 4359 hCoV-19/Canada/BC_22514413/2020\|EPI_ISL_462837\|202 | ···················· | ···················· | ····················· | ···················· | ···················· | ···················· |
| D | 4360 hCoV-19/Canada/BC_22946611/2020\|EPI_ISL_462838\|202 | ···················· | ···················· | ····················· | ···················· | ···················· | ···················· |
| D | 4361 hCoV-19/Canada/BC_23064916/2020\|EPI_ISL_462840\|202 | ···················· | ···················· | ····················· | ···················· | ···················· | ···················· |
| D | 4362 hCoV-19/Canada/BC_23094312/2020\|EPI_ISL_462841\|202 | ···················· | ···················· | ····················· | ···················· | ···················· | ···················· |
| D | 4363 hCoV-19/Canada/BC_23098110/2020\|EPI_ISL_462842\|202 | ···················· | ···················· | ····················· | ···················· | ···················· | ···················· |
| D | 4364 hCoV-19/Canada/BC_23108423/2020\|EPI_ISL_462843\|202 | ···················· | ···················· | ····················· | ···················· | ···················· | ···················· |
| D | 4365 hCoV-19/USA/MN-MDH-1000/2020\|EPI_ISL_462845\|2020-0 | ···················· | ···················· | ····················· | ···················· | ···················· | ···················· |
| D | 4366 hCoV-19/USA/MN-MDH-1001/2020\|EPI_ISL_462846\|2020-0 | ···················· | ···················· | ····················· | ···················· | ···················· | ···················· |
| D | 4367 hCoV-19/USA/MN-MDH-1002/2020\|EPI_ISL_462847\|2020-0 | ···················· | ···················· | ····················· | ···················· | ···················· | ···················· |
| D | 4368 hCoV-19/USA/MN-MDH-1003/2020\|EPI_ISL_462848\|2020-0 | ···················· | ···················· | ····················· | ···················· | ···················· | ···················· |
| D | 4369 hCoV-19/USA/MN-MDH-1004/2020\|EPI_ISL_462849\|2020-0 | ···················· | ···················· | ····················· | ···················· | ···················· | ···················· |
| D | 4370 hCoV-19/USA/MN-MDH-1005/2020\|EPI_ISL_462850\|2020-0 | ···················· | ···················· | ····················· | ···················· | ···················· | ···················· |
| D | 4371 hCoV-19/USA/MN-MDH-1006/2020\|EPI_ISL_462851\|2020-0 | ···················· | ···················· | ····················· | ···················· | ···················· | ···················· |
| D | 4372 hCoV-19/USA/MN-MDH-1007/2020\|EPI_ISL_462852\|2020-0 | ···················· | ···················· | ····················· | ···················· | ···················· | ···················· |
| D | 4373 hCoV-19/USA/MN-MDH-1008/2020\|EPI_ISL_462853\|2020-0 | ···················· | ···················· | ····················· | ···················· | ···················· | ···················· |
| D | 4374 hCoV-19/USA/MN-MDH-1009/2020\|EPI_ISL_462854\|2020-0 | ···················· | ···················· | ····················· | ···················· | ···················· | ···················· |
| D | 4375 hCoV-19/USA/MN-MDH-1010/2020\|EPI_ISL_462855\|2020-0 | ···················· | ···················· | ····················· | ···················· | ···················· | ···················· |
| D | 4376 hCoV-19/USA/MN-MDH-1011/2020\|EPI_ISL_462856\|2020-0 | ···················· | ···················· | ····················· | ···················· | ···················· | ···················· |
| D | 4377 hCoV-19/USA/MN-MDH-1012/2020\|EPI_ISL_462857\|2020-0 | ···················· | ···················· | ····················· | ···················· | ···················· | ···················· |
| D | 4378 hCoV-19/USA/MN-MDH-1013/2020\|EPI_ISL_462858\|2020-0 | ···················· | ···················· | ····················· | ···················· | ···················· | ···················· |
| D | 4379 hCoV-19/USA/MN-MDH-1014/2020\|EPI_ISL_462859\|2020-0 | ···················· | ···················· | ····················· | ···················· | ···················· | ···················· |
| D | 4380 hCoV-19/USA/MN-MDH-1015/2020\|EPI_ISL_462860\|2020-0 | ···················· | ···················· | ····················· | ···················· | ···················· | ···················· |
| D | 4381 hCoV-19/USA/MN-MDH-1016/2020\|EPI_ISL_462861\|2020-0 | ···················· | ···················· | ····················· | ···················· | ···················· | ···················· |
| D | 4382 hCoV-19/USA/MN-MDH-1017/2020\|EPI_ISL_462862\|2020-0 | ···················· | ···················· | ····················· | ···················· | ···················· | ···················· |
| D | 4383 hCoV-19/USA/MN-MDH-1019/2020\|EPI_ISL_462864\|2020-0 | ···················· | ···················· | ····················· | ···················· | ···················· | ···················· |
| D | 4384 hCoV-19/USA/MN-MDH-1020/2020\|EPI_ISL_462865\|2020-0 | ···················· | ·A·················· | ····················· | ···················· | ···················· | ···················· |
| D | 4385 hCoV-19/USA/MN-MDH-1021/2020\|EPI_ISL_462866\|2020-0 | ···················· | ·A·················· | ····················· | ···················· | ···················· | ···················· |
| D | 4386 hCoV-19/USA/MN-MDH-1022/2020\|EPI_ISL_462867\|2020-0 | ···················· | ·A·················· | ····················· | ···················· | ···················· | ···················· |
| D | 4387 hCoV-19/USA/MN-MDH-1023/2020\|EPI_ISL_462868\|2020-0 | ···················· | ·A·················· | ····················· | ···················· | ···················· | ···················· |

| | | | | | | | | |
|---|---|---|---|---|---|---|---|---|
| D | 4388 hCoV-19/USA/MN-MDH-1024/2020\|EPI_ISL_462869\|2020-0 | .................... | .A.................. | .................... | .................... | .................... | .................... | |
| D | 4389 hCoV-19/USA/MN-MDH-1025/2020\|EPI_ISL_462870\|2020-0 | .................... | .A.................. | .................... | .................... | .................... | .................... | |
| D | 4390 hCoV-19/USA/MN-MDH-1026/2020\|EPI_ISL_462871\|2020-0 | .................... | .A.................. | .................... | .................... | .................... | .................... | |
| D | 4391 hCoV-19/USA/MN-MDH-1027/2020\|EPI_ISL_462872\|2020-0 | .................... | .A.................. | .................... | .................... | .................... | .................... | |
| D | 4392 hCoV-19/USA/MN-MDH-1028/2020\|EPI_ISL_462873\|2020-0 | .................... | .A.................. | .................... | .................... | .................... | .................... | |
| D | 4393 hCoV-19/USA/MN-MDH-1029/2020\|EPI_ISL_462874\|2020-0 | .................... | .A.................. | .................... | .................... | .................... | .................... | |
| D | 4394 hCoV-19/USA/MN-MDH-1030/2020\|EPI_ISL_462875\|2020-0 | .................... | .A.................. | .................... | .................... | .................... | .................... | |
| D | 4395 hCoV-19/USA/MN-MDH-1031/2020\|EPI_ISL_462876\|2020-0 | .................... | .A.................. | .................... | .................... | .................... | .................... | |
| D | 4396 hCoV-19/USA/MN-MDH-1032/2020\|EPI_ISL_462877\|2020-0 | .................... | .A.................. | .................... | .................... | .................... | .................... | |
| D | 4397 hCoV-19/USA/MN-MDH-1033/2020\|EPI_ISL_462878\|2020-0 | .................... | .A.................. | .................... | .................... | .................... | .................... | |
| D | 4398 hCoV-19/USA/MN-MDH-1034/2020\|EPI_ISL_462879\|2020-0 | .................... | .A.................. | .................... | .................... | .................... | .................... | |
| D | 4399 hCoV-19/USA/MN-MDH-1035/2020\|EPI_ISL_462880\|2020-0 | .................... | .A.................. | .................... | .................... | .................... | .................... | |
| D | 4400 hCoV-19/USA/MN-MDH-1036/2020\|EPI_ISL_462881\|2020-0 | .................... | .A.................. | .................... | .................... | .................... | .................... | |
| D | 4401 hCoV-19/USA/MN-MDH-1037/2020\|EPI_ISL_462882\|2020-0 | .................... | .A.................. | .................... | .................... | .................... | .................... | |
| D | 4402 hCoV-19/USA/MN-MDH-1038/2020\|EPI_ISL_462883\|2020-0 | .................... | .A.................. | .................... | .................... | .................... | .................... | |
| D | 4403 hCoV-19/USA/MN-MDH-1039/2020\|EPI_ISL_462884\|2020-0 | .................... | .A.................. | .................... | .................... | .................... | .................... | |
| D | 4404 hCoV-19/USA/MN-MDH-1040/2020\|EPI_ISL_462885\|2020-0 | .................... | .A.................. | .................... | .................... | .................... | .................... | |
| D | 4405 hCoV-19/USA/MN-MDH-1041/2020\|EPI_ISL_462886\|2020-0 | .................... | .A.................. | .................... | .................... | .................... | .................... | |
| D | 4406 hCoV-19/USA/MN-MDH-1042/2020\|EPI_ISL_462887\|2020-0 | .................... | .................... | .................... | .................... | .................... | .................... | |
| D | 4407 hCoV-19/USA/MN-MDH-1043/2020\|EPI_ISL_462888\|2020-0 | .................... | .................... | .................... | .................... | .................... | .................... | |
| D | 4408 hCoV-19/USA/MN-MDH-1044/2020\|EPI_ISL_462889\|2020-0 | .................... | .................... | .................... | .................... | .................... | .................... | |
| D | 4409 hCoV-19/USA/MN-MDH-1045/2020\|EPI_ISL_462890\|2020-0 | .................... | .................... | .................... | .................... | .................... | .................... | |
| D | 4410 hCoV-19/USA/MN-MDH-1046/2020\|EPI_ISL_462891\|2020-0 | .................... | .A.................. | .................... | .................... | .................... | .................... | |
| D | 4411 hCoV-19/USA/MN-MDH-1047/2020\|EPI_ISL_462892\|2020-0 | .................... | .A.................. | .................... | .................... | .................... | .................... | |
| D | 4412 hCoV-19/USA/MN-MDH-1048/2020\|EPI_ISL_462893\|2020-0 | .................... | .A.................. | .................... | .................... | .................... | .................... | |
| D | 4413 hCoV-19/USA/MN-MDH-1049/2020\|EPI_ISL_462894\|2020-0 | .................... | .A.................. | .................... | .................... | .................... | .................... | |
| D | 4414 hCoV-19/USA/MN-MDH-1050/2020\|EPI_ISL_462895\|2020-0 | .................... | .A.................. | .................... | .................... | .................... | .................... | |
| D | 4415 hCoV-19/USA/MN-MDH-1052/2020\|EPI_ISL_462897\|2020-0 | .................... | .A.................. | .................... | .................... | .................... | .................... | |
| D | 4416 hCoV-19/USA/MN-MDH-1053/2020\|EPI_ISL_462898\|2020-0 | .................... | .A.................. | .................... | .................... | .................... | .................... | |
| D | 4417 hCoV-19/USA/MN-MDH-1054/2020\|EPI_ISL_462899\|2020-0 | .................... | .A.................. | .................... | .................... | .................... | .................... | |
| D | 4418 hCoV-19/England/20119011704/2020\|EPI_ISL_465030\|20 | .................... | .................... | .................... | .................... | .................... | .................... | |
| D | 4421 hCoV-19/England/20119023104/2020\|EPI_ISL_465034\|20 | .................... | .................... | .................... | .................... | .................... | ..N................. | |
| D | 4423 hCoV-19/England/20119027904/2020\|EPI_ISL_465038\|20 | .................... | .................... | .................... | .................... | .................... | .................... | |
| D | 4424 hCoV-19/England/20119029504/2020\|EPI_ISL_465039\|20 | .................... | .................... | .................... | .................... | .................... | .................... | |
| D | 4425 hCoV-19/England/20119029804/2020\|EPI_ISL_465040\|20 | .................... | .................... | .................... | .................... | .................... | .................... | |
| D | 4428 hCoV-19/England/20119031304/2020\|EPI_ISL_465043\|20 | .................... | .................... | .................... | .................... | .................... | .................... | |
| D | 4431 hCoV-19/England/20119032104/2020\|EPI_ISL_465046\|20 | .................... | .................... | .................... | .................... | .................... | .................... | |
| D | 4432 hCoV-19/England/20119033504/2020\|EPI_ISL_465049\|20 | .................... | .................... | .................... | .................... | .................... | .................... | |
| D | 4433 hCoV-19/England/20119034404/2020\|EPI_ISL_465050\|20 | .................... | .................... | .................... | .................... | .................... | .................... | |
| D | 4434 hCoV-19/England/20119037304/2020\|EPI_ISL_465052\|20 | .................... | .................... | .................... | .................... | .................... | .................... | |
| D | 4435 hCoV-19/England/20119039204/2020\|EPI_ISL_465054\|20 | .................... | .................... | .................... | .................... | .................... | .................... | |
| D | 4436 hCoV-19/England/20119040406/2020\|EPI_ISL_465057\|20 | .................... | .................... | .................... | .................... | .................... | .................... | |
| D | 4438 hCoV-19/England/20119041204/2020\|EPI_ISL_465059\|20 | .................... | .................... | .................... | .................... | .................... | .................... | |
| D | 4439 hCoV-19/England/20119041304/2020\|EPI_ISL_465060\|20 | .................... | .................... | .................... | .................... | .................... | .................... | |
| D | 4442 hCoV-19/England/20119044404/2020\|EPI_ISL_465065\|20 | .................... | .................... | .................... | .................... | .................... | .................... | |
| D | 4443 hCoV-19/England/20119047104/2020\|EPI_ISL_465068\|20 | .................... | .................... | .................... | .................... | .................... | .................... | |
| D | 4444 hCoV-19/England/201580203/2020\|EPI_ISL_466361\|2020 | .................... | .................... | .................... | .................... | .................... | .................... | |
| D | 4445 hCoV-19/England/201580278/2020\|EPI_ISL_466363\|2020 | .................... | .................... | .................... | .................... | .................... | .................... | |
| D | 4446 hCoV-19/England/201580284/2020\|EPI_ISL_466365\|2020 | .................... | .................... | .................... | .................... | .................... | .................... | |
| D | 4447 hCoV-19/England/201580285/2020\|EPI_ISL_466366\|2020 | .................... | .................... | .................... | .................... | .................... | .................... | |
| D | 4448 hCoV-19/England/201620024/2020\|EPI_ISL_466385\|2020 | .................... | .................... | .................... | .................... | .................... | .................... | |
| D | 4449 hCoV-19/England/20162003404/2020\|EPI_ISL_466389\|20 | .................... | .................... | .................... | .................... | ..NN................ | .................... | |
| D | 4450 hCoV-19/England/201620041/2020\|EPI_ISL_466391\|2020 | .................... | .................... | .................... | .................... | .................... | .................... | |
| D | 4451 hCoV-19/England/201620042/2020\|EPI_ISL_466392\|2020 | .................... | .................... | .................... | .................... | .................... | .................... | |
| D | 4452 hCoV-19/England/201620047/2020\|EPI_ISL_466393\|2020 | .................... | .................... | .................... | .................... | .................... | .................... | |
| D | 4453 hCoV-19/England/201620049/2020\|EPI_ISL_466395\|2020 | .................... | .................... | .................... | .................... | .................... | .................... | |
| D | 4454 hCoV-19/England/20162005404/2020\|EPI_ISL_466399\|20 | .................... | .................... | .................... | .................... | ......NN............ | .................... | |
| D | 4455 hCoV-19/England/BRIS-1311C6/2020\|EPI_ISL_467695\|20 | .................... | .................... | .................... | .................... | .................... | .................... | |
| D | 4456 hCoV-19/England/BRIS-13122D/2020\|EPI_ISL_467699\|20 | .................... | .................... | .................... | .................... | .................... | .................... | |
| D | 4457 hCoV-19/England/NORT-282D0D/2020\|EPI_ISL_472228\|20 | .................... | .................... | .................... | .................... | .................... | .................... | |
| D | 4458 hCoV-19/England/CAMB-1B4B63/2020\|EPI_ISL_473500\|20 | .................... | .................... | .................... | .................... | .................... | .................... | |
| D | 4459 hCoV-19/England/CAMB-1B4BBE/2020\|EPI_ISL_473501\|20 | .................... | .................... | .................... | .................... | .................... | .................... | |
| D | 4460 hCoV-19/England/CAMB-1B4C15/2020\|EPI_ISL_473503\|20 | .................... | .................... | .................... | .................... | .................... | .................... | |
| D | 4461 hCoV-19/England/CAMB-1B4C42/2020\|EPI_ISL_473504\|20 | .................... | .................... | .................... | .................... | .................... | .................... | |
| D | 4462 hCoV-19/England/CAMB-1B4D03/2020\|EPI_ISL_473507\|20 | .................... | .................... | .................... | .................... | .................... | .................... | |
| D | 4463 hCoV-19/Scotland/CVR2046/2020\|EPI_ISL_473509\|2020- | .................... | .................... | .................... | .................... | .................... | .................... | |
| D | 4464 hCoV-19/Scotland/CVR2088/2020\|EPI_ISL_473510\|2020- | .................... | .................... | .................... | .................... | .................... | .................... | |
| D | 4465 hCoV-19/Scotland/CVR2163/2020\|EPI_ISL_473511\|2020- | .................... | .................... | .................... | .................... | .................... | .................... | |
| D | 4466 hCoV-19/Scotland/CVR2264/2020\|EPI_ISL_473512\|2020- | .................... | .................... | .................... | .................... | .................... | .................... | |
| D | 4467 hCoV-19/Scotland/CVR2537/2020\|EPI_ISL_473513\|2020- | .................... | .................... | .................... | .................... | .................... | .................... | |
| D | 4468 hCoV-19/Scotland/CVR2550/2020\|EPI_ISL_473514\|2020- | .................... | .................... | .................... | .................... | .................... | .................... | |
| D | 4469 hCoV-19/Scotland/CVR2556/2020\|EPI_ISL_473515\|2020- | .................... | .................... | .................... | .................... | .................... | .................... | |
| D | 4470 hCoV-19/Scotland/CVR2572/2020\|EPI_ISL_473516\|2020- | .................... | .................... | .................... | .................... | .................... | .................... | |
| D | 4471 hCoV-19/Scotland/CVR2579/2020\|EPI_ISL_473517\|2020- | .................... | ...................A | .................... | .................... | .................... | .................... | 1 |
| D | 4472 hCoV-19/Scotland/CVR2580/2020\|EPI_ISL_473518\|2020- | .................... | .................... | .................... | .................... | .................... | .................... | |
| D | 4473 hCoV-19/Scotland/CVR2583/2020\|EPI_ISL_473519\|2020- | .................... | ...................A | .................... | .................... | .................... | .................... | 1 |
| D | 4474 hCoV-19/Scotland/CVR2586/2020\|EPI_ISL_473520\|2020- | .................... | .................... | .................... | .................... | .................... | .................... | |
| D | 4475 hCoV-19/Scotland/CVR2587/2020\|EPI_ISL_473521\|2020- | .................... | .................... | .................... | .................... | .................... | .................... | |

| | | | | | | | | | |
|---|---|---|---|---|---|---|---|---|---|
| D | 4476 | hCoV-19/Scotland/CVR2588/2020\|EPI_ISL_473522\|2020- | ···················· | ···················· | ···················· | ···················· | ···················· | ···················· | |
| D | 4478 | hCoV-19/Scotland/CVR2605/2020\|EPI_ISL_473526\|2020- | ···················· | ·A·················· | ···················· | ···················· | ···················· | ···················· | |
| D | 4479 | hCoV-19/Scotland/CVR2606/2020\|EPI_ISL_473527\|2020- | ···················· | ···················· | ···················· | ···················· | ···················· | ···················· | |
| D | 4480 | hCoV-19/Scotland/CVR2647/2020\|EPI_ISL_473528\|2020- | ···················· | ···················· | ···················· | ···················· | ···················· | ···················· | |
| D | 4481 | hCoV-19/Scotland/CVR2670/2020\|EPI_ISL_473529\|2020- | ···················· | ···················· | ···················· | ···················· | ···················· | ···················· | |
| D | 4482 | hCoV-19/Scotland/CVR2703/2020\|EPI_ISL_473532\|2020- | ···················· | ···················· | ···················· | ···················· | ···················· | ···················· | |
| D | 4483 | hCoV-19/Scotland/CVR2710/2020\|EPI_ISL_473533\|2020- | ···················· | ···················· | ···················· | ···················· | ···················· | ···················· | |
| D | 4484 | hCoV-19/Scotland/CVR2713/2020\|EPI_ISL_473534\|2020- | ···················· | ···················A | ···················· | ···················· | ···················· | ···················· | 1 |
| D | 4485 | hCoV-19/Scotland/CVR2714/2020\|EPI_ISL_473535\|2020- | ···················· | ···················· | ···················· | ···················· | ···················· | ···················· | |
| D | 4486 | hCoV-19/Scotland/CVR2715/2020\|EPI_ISL_473536\|2020- | ···················· | ···················A | ···················· | ···················· | ···················· | ···················· | 1 |
| D | 4487 | hCoV-19/Scotland/CVR2716/2020\|EPI_ISL_473537\|2020- | ···················· | ···················· | ···················· | ···················· | ···················· | ···················· | |
| D | 4488 | hCoV-19/Scotland/CVR2719/2020\|EPI_ISL_473540\|2020- | ···················· | ···················· | ···················· | ···················· | ···················· | ···················· | |
| D | 4489 | hCoV-19/Scotland/CVR2720/2020\|EPI_ISL_473541\|2020- | ···················· | ···················· | ···················· | ···················· | ···················· | ···················· | |
| D | 4490 | hCoV-19/Scotland/CVR2722/2020\|EPI_ISL_473543\|2020- | ···················· | ···················· | ···················· | ···················· | ···················· | ···················· | |
| D | 4491 | hCoV-19/Scotland/CVR2723/2020\|EPI_ISL_473544\|2020- | ···················· | ···················A | ···················· | ···················· | ···················· | ···················· | 1 |
| D | 4492 | hCoV-19/Scotland/CVR2725/2020\|EPI_ISL_473546\|2020- | ···················· | ···················A | ···················· | ···················· | ···················· | ···················· | 1 |
| D | 4493 | hCoV-19/Scotland/CVR2726/2020\|EPI_ISL_473547\|2020- | ···················· | ···················· | ···················· | ···················· | ···················· | ···················· | |
| D | 4494 | hCoV-19/Scotland/CVR2727/2020\|EPI_ISL_473548\|2020- | ···················· | ·A·················· | ···················· | ···················· | ···················· | ···················· | |
| D | 4495 | hCoV-19/Scotland/CVR2729/2020\|EPI_ISL_473549\|2020- | ···················· | ···················· | ···················· | ···················· | ···················· | ···················· | |
| D | 4496 | hCoV-19/Scotland/CVR2730/2020\|EPI_ISL_473550\|2020- | ···················· | ···················· | ···················· | ···················· | ···················· | ···················· | |
| D | 4497 | hCoV-19/Scotland/CVR2731/2020\|EPI_ISL_473551\|2020- | ···················· | ···················· | ···················· | ···················· | ···················· | ···················· | |
| D | 4498 | hCoV-19/Scotland/CVR2760/2020\|EPI_ISL_473552\|2020- | ···················· | ··············C····· | ···················· | ···················· | ···················· | ···················· | |
| D | 4499 | hCoV-19/Scotland/CVR2761/2020\|EPI_ISL_473553\|2020- | ···················· | ···················· | ···················· | ···················· | ···················· | ···················· | |
| D | 4500 | hCoV-19/Scotland/CVR2762/2020\|EPI_ISL_473554\|2020- | ···················· | ···················· | ···················· | ···················· | ···················· | ···················· | |
| D | 4502 | hCoV-19/Scotland/CVR2769/2020\|EPI_ISL_473557\|2020- | ···················· | ···················· | ···················· | ···················· | ···················· | ···················· | |
| D | 4503 | hCoV-19/Scotland/CVR2772/2020\|EPI_ISL_473558\|2020- | ···················· | ···················· | ···················· | ···················· | ···················· | ···················· | |
| D | 4505 | hCoV-19/Scotland/CVR2778/2020\|EPI_ISL_473560\|2020- | ···················· | ·A·················· | ···················· | ···················· | ···················· | ···················· | |
| D | 4506 | hCoV-19/Scotland/CVR2779/2020\|EPI_ISL_473561\|2020- | ···················· | ·A·················· | ···················· | ···················· | ···················· | ···················· | |
| D | 4507 | hCoV-19/Scotland/CVR2781/2020\|EPI_ISL_473562\|2020- | ···················· | ···················· | ···················· | ···················· | ···················· | ···················· | |
| D | 4510 | hCoV-19/Spain/CM-IBV-003777/2020\|EPI_ISL_474810\|20 | ···················· | ···················· | ···················· | ···················· | ···················· | ···················· | |
| D | 4516 | hCoV-19/Spain/CM-IBV-003792/2020\|EPI_ISL_474818\|20 | ···················· | ···················· | ···················· | ···················· | ···················· | ···················· | |
| D | 4531 | hCoV-19/Spain/AN-IBV-002998/2020\|EPI_ISL_474836\|20 | ···················· | ···················· | ···················· | ···················· | ···················· | ···················· | |
| D | 4537 | hCoV-19/Spain/AN-IBV-002921/2020\|EPI_ISL_474845\|20 | ···················· | ···················· | ···················· | ···················· | ···················· | ···················· | |
| D | 4546 | hCoV-19/Spain/AN-IBV-002875/2020\|EPI_ISL_474862\|20 | ···················· | ···················· | ···················· | ···················· | ··N················· | ···················· | |
| D | 4549 | hCoV-19/Spain/AN-IBV-002891/2020\|EPI_ISL_474868\|20 | ···················· | ···················· | ···················· | ···················· | ···················· | ···················· | |
| D | 4550 | hCoV-19/Spain/AN-IBV-002908/2020\|EPI_ISL_474869\|20 | ···················· | ···················· | ···················· | ···················· | ···················· | ···················· | |
| D | 4551 | hCoV-19/Spain/AN-IBV-002915/2020\|EPI_ISL_474870\|20 | ···················· | ···················· | ···················· | ···················· | ···················· | ···················· | |
| D | 4552 | hCoV-19/Spain/AN-IBV-002916/2020\|EPI_ISL_474871\|20 | ···················· | ···················· | ···················· | ···················· | ···N·NN············· | ···················· | 1 |
| D | 4555 | hCoV-19/Spain/AN-IBV-002930/2020\|EPI_ISL_474874\|20 | ···················· | ···················· | ···················· | ···················· | ···················· | ···················· | |
| D | 4559 | hCoV-19/Spain/AN-IBV-002958/2020\|EPI_ISL_474880\|20 | ···················· | ···················· | ···················· | ···················· | ···················· | ···················· | |
| D | 4565 | hCoV-19/Spain/AN-IBV-002990/2020\|EPI_ISL_474888\|20 | ···················· | ···················· | ···················· | ···················· | ···················· | ···················· | |
| D | 4567 | hCoV-19/Spain/AN-IBV-003010/2020\|EPI_ISL_474890\|20 | ···················· | ···················· | ···················· | ···················· | ···················· | ···················· | |
| D | 4569 | hCoV-19/Spain/AN-IBV-003020/2020\|EPI_ISL_474892\|20 | ···················· | ···················· | ···················· | ···················· | ···················· | ···················· | |
| D | 4570 | hCoV-19/Canada/BC_04701167/2020\|EPI_ISL_477050\|202 | ···················· | ···················· | ···················· | ···················· | ···················· | ···················· | |
| D | 4571 | hCoV-19/Canada/BC_04709878/2020\|EPI_ISL_477051\|202 | ···················· | ···················· | ···················· | ···················· | ···················· | ···················· | |
| D | 4572 | hCoV-19/Canada/BC_04710049/2020\|EPI_ISL_477052\|202 | ···················· | ···················· | ···················· | ···················· | ···················· | ···················· | |
| D | 4573 | hCoV-19/Canada/BC_04806249/2020\|EPI_ISL_477054\|202 | ···················· | ···················· | ···················· | ···················· | ···················· | ···················· | |
| D | 4574 | hCoV-19/Canada/BC_04806819/2020\|EPI_ISL_477056\|202 | ···················· | ···················· | ···················· | ···················· | ···················· | ···················· | |
| D | 4575 | hCoV-19/Canada/BC_04865055/2020\|EPI_ISL_477057\|202 | ···················· | ···················· | ···················· | ···················· | ···················· | ···················· | |
| D | 4576 | hCoV-19/Canada/BC_06074915/2020\|EPI_ISL_477058\|202 | ···················· | ···················· | ···················· | ···················· | ···················· | ···················· | |
| D | 4577 | hCoV-19/Canada/BC_06514625/2020\|EPI_ISL_477059\|202 | ···················· | ···················· | ···················· | ···················· | ···················· | ···················· | |
| D | 4578 | hCoV-19/Canada/BC_07353444/2020\|EPI_ISL_477060\|202 | ···················· | ···················· | ···················· | ···················· | ···················· | ···················· | |
| D | 4579 | hCoV-19/Canada/BC_07653696/2020\|EPI_ISL_477061\|202 | ···················· | ···················· | ···················· | ···················· | ···················· | ···················· | |
| D | 4580 | hCoV-19/Canada/BC_08164457/2020\|EPI_ISL_477062\|202 | ···················· | ···················· | ···················· | ···················· | ···················· | ···················· | |
| D | 4581 | hCoV-19/Canada/BC_08312782/2020\|EPI_ISL_477063\|202 | ···················· | ···················· | ···················· | ···················· | ···················· | ···················· | |
| D | 4582 | hCoV-19/Canada/BC_08534834/2020\|EPI_ISL_477064\|202 | ···················· | ···················· | ···················· | ···················· | ···················· | ···················· | |
| D | 4583 | hCoV-19/Canada/BC_08855516/2020\|EPI_ISL_477065\|202 | ···················· | ···················· | ···················· | ···················· | ···················· | ···················· | |
| D | 4584 | hCoV-19/Canada/BC_08859333/2020\|EPI_ISL_477066\|202 | ···················· | ···················· | ···················· | ···················· | ···················· | ···················· | |
| D | 4585 | hCoV-19/Canada/BC_08865483/2020\|EPI_ISL_477067\|202 | ···················· | ···················· | ···················· | ···················· | ···················· | ···················· | |
| D | 4586 | hCoV-19/Canada/BC_08984631/2020\|EPI_ISL_477068\|202 | ···················· | ···················· | ···················· | ···················· | ···················· | ···················· | |
| D | 4587 | hCoV-19/Canada/BC_08987762/2020\|EPI_ISL_477069\|202 | ···················· | ···················· | ···················· | ···················· | ···················· | ···················· | |
| D | 4588 | hCoV-19/Canada/BC_09273580/2020\|EPI_ISL_477070\|202 | ···················· | ···················· | ···················· | ···················· | ···················· | ···················· | |
| D | 4589 | hCoV-19/Canada/BC_09625012/2020\|EPI_ISL_477071\|202 | ···················· | ···················· | ···················· | ···················· | ···················· | ···················· | |
| D | 4590 | hCoV-19/Canada/BC_09819716/2020\|EPI_ISL_477072\|202 | ···················· | ···················· | ···················· | ···················· | ···················· | ···················· | |
| D | 4591 | hCoV-19/Canada/BC_09963377/2020\|EPI_ISL_477073\|202 | ···················· | ···················· | ···················· | ···················· | ···················· | ···················· | |
| D | 4592 | hCoV-19/Canada/BC_09965869/2020\|EPI_ISL_477074\|202 | ···················· | ···················· | ···················· | ···················· | ···················· | ···················· | |
| D | 4593 | hCoV-19/Canada/BC_09986014/2020\|EPI_ISL_477075\|202 | ···················· | ···················· | ···················· | ···················· | ···················· | ···················· | |
| D | 4594 | hCoV-19/Canada/BC_19879538/2020\|EPI_ISL_477076\|202 | ···················· | ···················· | ···················· | ···················· | ···················· | ···················· | |
| D | 4595 | hCoV-19/Canada/BC_10404688/2020\|EPI_ISL_477079\|202 | ···················· | ···················· | ···················· | ···················· | ···················· | ···················· | |
| D | 4596 | hCoV-19/Canada/BC_03990815/2020\|EPI_ISL_477090\|202 | ···················· | ···················· | ···················· | ···················· | ···················· | ···················· | |
| D | 4597 | hCoV-19/Canada/BC_03998412/2020\|EPI_ISL_477091\|202 | ···················· | ···················· | ···················· | ···················· | ···················· | ···················· | |
| D | 4598 | hCoV-19/Canada/BC_04000917/2020\|EPI_ISL_477092\|202 | ···················· | ···················· | ···················· | ···················· | ···················· | ···················· | |
| D | 4599 | hCoV-19/Canada/BC_07905258/2020\|EPI_ISL_477093\|202 | ···················· | ···················· | ···················· | ···················· | ···················· | ···················· | |
| D | 4600 | hCoV-19/Canada/BC_07905790/2020\|EPI_ISL_477094\|202 | ···················· | ···················· | ···················· | ···················· | ···················· | ···················· | |
| D | 4601 | hCoV-19/Canada/BC_07911261/2020\|EPI_ISL_477095\|202 | ···················· | ···················· | ···················· | ···················· | ···················· | ···················· | |
| D | 4602 | hCoV-19/Canada/BC_07912118/2020\|EPI_ISL_477096\|202 | ···················· | ···················· | ···················· | ···················· | ···················· | ···················· | |
| D | 4603 | hCoV-19/Canada/BC_07916655/2020\|EPI_ISL_477098\|202 | ···················· | ···················· | ···················· | ···················· | ···················· | ···················· | |
| D | 4604 | hCoV-19/Canada/BC_07917243/2020\|EPI_ISL_477099\|202 | ···················· | ···················· | ···················· | ···················· | ···················· | ···················· | |

| | | | | | | | | |
|---|---|---|---|---|---|---|---|---|
| D | 4605 | hCoV-19/England/LIVE-A0C2C/2020\|EPI_ISL_478392\|202 | ···················· | ···················· | ····················· | ···················· | ···················· | ···················· |
| D | 4606 | hCoV-19/England/LIVE-A0C68/2020\|EPI_ISL_478394\|202 | ···················· | ···················· | ····················· | ···················· | ···················· | ···················· |
| D | 4607 | hCoV-19/England/LIVE-A0C95/2020\|EPI_ISL_478395\|202 | ···················· | ···················· | ····················· | ···················· | ···················· | ···················· |
| D | 4608 | hCoV-19/England/LIVE-A0D38/2020\|EPI_ISL_478396\|202 | ···················· | ···················· | ····················· | ···················· | ···················· | ···················· |
| D | 4609 | hCoV-19/Canada/BC_21228218/2020\|EPI_ISL_462802\|202 | ···················· | ···················· | ····················· | ···················· | ···················· | ···················· |
| D | 4610 | hCoV-19/Canada/BC_21317517/2020\|EPI_ISL_462805\|202 | ···················· | ···················· | ····················· | ···················· | ···················· | ···················· |
| D | 4611 | hCoV-19/Canada/BC_21333019/2020\|EPI_ISL_462806\|202 | ···················· | ···················· | ····················· | ···················· | ···················· | ···················· |
| D | 4612 | hCoV-19/Canada/BC_21346429/2020\|EPI_ISL_462807\|202 | ···················· | ···················· | ····················· | ···················· | ···················· | ···················· |
| D | 4613 | hCoV-19/Canada/BC_21350918/2020\|EPI_ISL_462808\|202 | ···················· | ···················· | ····················· | ···················· | ···················· | ···················· |
| D | 4614 | hCoV-19/Canada/BC_21368114/2020\|EPI_ISL_462809\|202 | ···················· | ···················· | ····················· | ···················· | ···················· | ···················· |
| D | 4615 | hCoV-19/Canada/BC_21385821/2020\|EPI_ISL_462810\|202 | ···················· | ···················· | ····················· | ···················· | ···················· | ···················· |
| D | 4616 | hCoV-19/Canada/BC_21388915/2020\|EPI_ISL_462811\|202 | ···················· | ···················· | ····················· | ···················· | ···················· | ···················· |
| D | 4617 | hCoV-19/Canada/BC_21426715/2020\|EPI_ISL_462812\|202 | ···················· | ···················· | ····················· | ···················· | ···················· | ···················· |
| D | 4618 | hCoV-19/Canada/BC_21446719/2020\|EPI_ISL_462813\|202 | ···················· | ···················· | ····················· | ···················· | ···················· | ···················· |
| D | 4619 | hCoV-19/Canada/BC_21461212/2020\|EPI_ISL_462814\|202 | ···················· | ···················· | ····················· | ···················· | ···················· | ···················· |
| D | 4620 | hCoV-19/Canada/BC_21462515/2020\|EPI_ISL_462815\|202 | ···················· | ···················· | ····················· | ···················· | ···················· | ···················· |
| D | 4621 | hCoV-19/Canada/BC_21466919/2020\|EPI_ISL_462816\|202 | ···················· | ···················· | ····················· | ···················· | ···················· | ···················· |
| D | 4622 | hCoV-19/Canada/BC_21480617/2020\|EPI_ISL_462818\|202 | ···················· | ···················· | ····················· | ···················· | ···················· | ···················· |
| D | 4623 | hCoV-19/Canada/BC_21927217/2020\|EPI_ISL_462819\|202 | ···················· | ···················· | ····················· | ···················· | ···················· | ···················· |
| D | 4624 | hCoV-19/Canada/BC_21993212/2020\|EPI_ISL_462820\|202 | ···················· | ···················· | ····················· | ···················· | ···················· | ···················· |
| D | 4625 | hCoV-19/Canada/BC_22032711/2020\|EPI_ISL_462821\|202 | ···················· | ···················· | ····················· | ···················· | ···················· | ···················· |
| D | 4626 | hCoV-19/Canada/BC_22037912/2020\|EPI_ISL_462822\|202 | ···················· | ···················· | ····················· | ···················· | ···················· | ···················· |
| D | 4627 | hCoV-19/Canada/BC_22070527/2020\|EPI_ISL_462824\|202 | ···················· | ···················· | ····················· | ···················· | ···················· | ···················· |
| D | 4628 | hCoV-19/Canada/BC_22156718/2020\|EPI_ISL_462825\|202 | ···················· | ···················· | ····················· | ···················· | ···················· | ···················· |
| D | 4629 | hCoV-19/Canada/BC_22218811/2020\|EPI_ISL_462826\|202 | ···················· | ···················· | ····················· | ···················· | ···················· | ···················· |
| D | 4630 | hCoV-19/Canada/BC_22230811/2020\|EPI_ISL_462827\|202 | ···················· | ···················· | ····················· | ···················· | ···················· | ···················· |
| D | 4631 | hCoV-19/Canada/BC_22332113/2020\|EPI_ISL_462828\|202 | ···················· | ···················· | ····················· | ···················· | ···················· | ···················· |
| D | 4632 | hCoV-19/Canada/BC_22349316/2020\|EPI_ISL_462829\|202 | ···················· | ···················· | ····················· | ···················· | ···················· | ···················· |
| D | 4633 | hCoV-19/India/OR-ILSCV11165/2020\|EPI_ISL_463010\|20 | ···················· | ···················· | ····················· | ···················· | ···················· | ···················· |
| D | 4634 | hCoV-19/India/OR-ILSCV12742/2020\|EPI_ISL_463013\|20 | ···················· | ···················· | ····················· | ···················· | ···················· | ···················· |
| D | 4635 | hCoV-19/India/OR-ILSCV12743/2020\|EPI_ISL_463014\|20 | ···················· | ···················· | ····················· | ···················· | ···················· | ···················· |
| D | 4636 | hCoV-19/India/OR-ILSCV12752/2020\|EPI_ISL_463017\|20 | ···················· | ···················· | ····················· | ···················· | ···················· | ···················· |
| D | 4637 | hCoV-19/India/OR-ILSCV12762/2020\|EPI_ISL_463018\|20 | ···················· | ···················· | ····················· | ···················· | ···················· | ···················· |
| D | 4638 | hCoV-19/India/OR-ILSCV13676/2020\|EPI_ISL_463019\|20 | ···················· | ···················· | ····················· | ···················· | ···················· | ···················· |
| D | 4639 | hCoV-19/India/OR-ILSCV13678/2020\|EPI_ISL_463020\|20 | ···················· | ···················· | ····················· | ···················· | ···················· | ···················· |
| D | 4640 | hCoV-19/India/OR-ILSCV13715/2020\|EPI_ISL_463022\|20 | ···················· | ···················· | ····················· | ···················· | ···················· | ···················· |
| D | 4641 | hCoV-19/India/OR-ILSCV13744/2020\|EPI_ISL_463023\|20 | ···················· | ···················· | ····················· | ···················· | ···················· | ···················· |
| D | 4642 | hCoV-19/India/OR-ILSCV13907/2020\|EPI_ISL_463024\|20 | ···················· | ···················· | ····················· | ···················· | ···················· | ···················· |
| D | 4643 | hCoV-19/India/OR-ILSCV13921/2020\|EPI_ISL_463025\|20 | ···················· | ···················· | ····················· | ···················· | ···················· | ···················· |
| D | 4644 | hCoV-19/India/OR-ILSCV13929/2020\|EPI_ISL_463026\|20 | ···················· | ···················· | ····················· | ···················· | ···················· | ···················· |
| D | 4645 | hCoV-19/India/OR-ILSCV14004/2020\|EPI_ISL_463027\|20 | ···················· | ···················· | ····················· | ···················· | ···················· | ···················· |
| D | 4646 | hCoV-19/India/OR-ILSCV14262/2020\|EPI_ISL_463029\|20 | ···················· | ···················· | ····················· | ···················· | ···················· | ···················· |
| D | 4647 | hCoV-19/India/OR-ILSCV14374/2020\|EPI_ISL_463031\|20 | ···················· | ···················· | ····················· | ···················· | ···················· | ···················· |
| D | 4648 | hCoV-19/India/OR-ILSCV14743/2020\|EPI_ISL_463032\|20 | ···················· | ···················· | ····················· | ···················· | ···················· | ···················· |
| D | 4649 | hCoV-19/India/OR-ILSCV14831/2020\|EPI_ISL_463033\|20 | ···················· | ···················· | ····················· | ···················· | ···················· | ···················· |
| D | 4650 | hCoV-19/India/OR-ILSCV14847/2020\|EPI_ISL_463034\|20 | ···················· | ···················· | ····················· | ···················· | ···················· | ···················· |
| D | 4651 | hCoV-19/India/OR-ILSCV15547/2020\|EPI_ISL_463036\|20 | ···················· | ···················· | ····················· | ···················· | ···················· | ···················· |
| D | 4652 | hCoV-19/India/OR-ILSCV15552/2020\|EPI_ISL_463037\|20 | ···················· | ···················· | ····················· | ···················· | ···················· | ···················· |
| D | 4653 | hCoV-19/India/OR-ILSCV15907/2020\|EPI_ISL_463038\|20 | ···················· | ···················· | ····················· | ···················· | ···················· | ···················· |
| D | 4654 | hCoV-19/India/OR-ILSCV15917/2020\|EPI_ISL_463039\|20 | ···················· | ···················· | ····················· | ···················· | ···················· | ···················· |
| D | 4656 | hCoV-19/India/OR-ILSCV16012/2020\|EPI_ISL_463043\|20 | ···················· | ···················· | ····················· | ···················· | ···················· | ···················· |
| D | 4658 | hCoV-19/England/201040706/2020\|EPI_ISL_464343\|2020 | ···················· | ···················· | ····················· | ···················· | ···················· | ···················· |
| D | 4659 | hCoV-19/England/201040707/2020\|EPI_ISL_464344\|2020 | ···················· | ···················· | ····················· | ···················· | ···················· | ···················· |
| D | 4660 | hCoV-19/England/201041387/2020\|EPI_ISL_464347\|2020 | ···················· | ···················· | ····················· | ···················· | ···················· | ···················· |
| D | 4662 | hCoV-19/England/201041493/2020\|EPI_ISL_464350\|2020 | ···················· | ···················· | ····················· | ···················· | ···················· | ···················· |
| D | 4663 | hCoV-19/England/201060003/2020\|EPI_ISL_464355\|2020 | ···················· | ···················· | ····················· | ···················· | ···················· | ···················· |
| D | 4664 | hCoV-19/England/201060004/2020\|EPI_ISL_464356\|2020 | ···················· | ···················· | ····················· | ···················· | ···················· | ···················· |
| D | 4665 | hCoV-19/England/201060007/2020\|EPI_ISL_464357\|2020 | ···················· | ···················· | ····················· | ···················· | ···················· | ···················· |
| D | 4666 | hCoV-19/England/201060008/2020\|EPI_ISL_464358\|2020 | ···················· | ···················· | ····················· | ···················· | ··N················· | ···················· |
| D | 4668 | hCoV-19/England/201060015/2020\|EPI_ISL_464361\|2020 | ···················· | ···················· | ····················· | ···················· | ···················· | ···················· |
| D | 4669 | hCoV-19/England/201060017/2020\|EPI_ISL_464362\|2020 | ···················· | ···················· | ····················· | ···················· | ···················· | ···················· |
| D | 4670 | hCoV-19/England/201060018/2020\|EPI_ISL_464363\|2020 | ···················· | ···················· | ····················· | ···················· | ···················· | ···················· |
| D | 4671 | hCoV-19/England/201060019/2020\|EPI_ISL_464364\|2020 | ···················· | ···················· | ····················· | ···················· | ···················· | ···N················ |
| D | 4672 | hCoV-19/England/201060020/2020\|EPI_ISL_464365\|2020 | ···················· | ···················· | ····················· | ···················· | ···················· | ···················· |
| D | 4673 | hCoV-19/England/201060023/2020\|EPI_ISL_464367\|2020 | ···················· | ···················· | ····················· | ···················· | ···················· | ···················· |
| D | 4675 | hCoV-19/England/201060026/2020\|EPI_ISL_464369\|2020 | ···················· | ···················· | ····················· | ···················· | ···················· | ···················· |
| D | 4678 | hCoV-19/England/201060035/2020\|EPI_ISL_464372\|2020 | ···················· | ···················· | ····················· | ···················· | ···················· | ···················· |
| D | 4680 | hCoV-19/England/201060038/2020\|EPI_ISL_464375\|2020 | ···················· | ···················· | ····················· | ···················· | ···················· | ···················· |
| D | 4681 | hCoV-19/England/201060041/2020\|EPI_ISL_464377\|2020 | ···················· | ···················· | ····················· | ···················· | ···T················ | ···················· |
| D | 4682 | hCoV-19/England/20119009904/2020\|EPI_ISL_465028\|20 | ···················· | ···················· | ····················· | ···················· | ···················· | ···················· |
| D | 4683 | hCoV-19/England/20119011604/2020\|EPI_ISL_465029\|20 | ···················· | ···················· | ····················· | ···················· | ···················· | ···················· |
| D | 4684 | hCoV-19/England/20119026204/2020\|EPI_ISL_465036\|20 | ···················· | ···················· | ····················· | ···················· | ···················· | ···················· |
| D | 4685 | hCoV-19/England/20119027704/2020\|EPI_ISL_465037\|20 | ···················· | ···················· | ····················· | ···················· | ················N··· | ···················· |
| D | 4686 | hCoV-19/England/20119047904/2020\|EPI_ISL_465070\|20 | ···················· | ···················· | ····················· | ···················· | ···················· | ···················· |
| D | 4688 | hCoV-19/England/20119049904/2020\|EPI_ISL_465073\|20 | ···················· | ·R·················· | ····················· | ···················· | ···················· | ···················· |
| D | 4689 | hCoV-19/England/20119054604/2020\|EPI_ISL_465075\|20 | ···················· | ···················· | ····················· | ···················· | ···················· | ···················· |
| D | 4690 | hCoV-19/England/20119056704/2020\|EPI_ISL_465078\|20 | ···················· | ···················· | ····················· | ···················· | ···················· | ···················· |
| D | 4691 | hCoV-19/England/20119057204/2020\|EPI_ISL_465079\|20 | ···················· | ···················· | ····················· | ···················· | ···················· | ···················· |

| | | | | | | | | |
|---|---|---|---|---|---|---|---|---|
| D | 4694 | hCoV-19/England/20119059404/2020\|EPI_ISL_465083\|20 | ···················· | ···················· | ···················· | ···················· | ···················· | ···················· |
| D | 4695 | hCoV-19/England/20119060904/2020\|EPI_ISL_465084\|20 | ···················· | ···················· | ···················· | ···················· | ···················· | ···················· |
| D | 4696 | hCoV-19/England/20119062704/2020\|EPI_ISL_465085\|20 | ···················· | ···················· | ···················· | ···················· | ···················· | ···················· |
| D | 4697 | hCoV-19/England/20119063104/2020\|EPI_ISL_465086\|20 | ···················· | ···················· | ···················· | ···················· | ···················· | ···················· |
| D | 4698 | hCoV-19/England/20119063404/2020\|EPI_ISL_465087\|20 | ···················· | ···················· | ···················· | ···················· | ···················· | ···················· |
| D | 4699 | hCoV-19/England/20119064904/2020\|EPI_ISL_465091\|20 | ···················· | ···················· | ···················· | ···················· | ···················· | ···················· |
| D | 4700 | hCoV-19/England/20119066104/2020\|EPI_ISL_465092\|20 | ···················· | ···················· | ···················· | ···················· | ···················· | ···················· |
| D | 4703 | hCoV-19/England/20119072704/2020\|EPI_ISL_465099\|20 | ···················· | ···················· | ···················· | ···················· | ···················· | ···················· |
| D | 4704 | hCoV-19/England/20212010104/2020\|EPI_ISL_465673\|20 | ···················· | ···················· | ···················· | ···················· | ···················· | ···················· |
| D | 4706 | hCoV-19/Canada/Qc-L00240569/2020\|EPI_ISL_465679\|20 | ···················· | ···················· | ···················· | ···················· | ···················· | ···················· |
| D | 4707 | hCoV-19/Canada/Qc-L00240594/2020\|EPI_ISL_465680\|20 | ···················· | ···················· | ···················· | ···················· | ···················· | ···················· |
| D | 4708 | hCoV-19/Canada/Qc-L00240642/2020\|EPI_ISL_465681\|20 | ···················· | ···················· | ···················· | ···················· | ···················· | ···················· |
| D | 4709 | hCoV-19/Canada/Qc-L00240666/2020\|EPI_ISL_465682\|20 | ···················· | ···················· | ···················· | ···················· | ···················· | ···················· |
| D | 4710 | hCoV-19/Canada/Qc-L00240746/2020\|EPI_ISL_465683\|20 | ···················· | ···················· | ···················· | ···················· | ···················· | ···················· |
| D | 4711 | hCoV-19/Canada/Qc-L00240750/2020\|EPI_ISL_465684\|20 | ···················· | ···················· | ···················· | ···················· | ···················· | ···················· |
| D | 4712 | hCoV-19/Canada/Qc-L00240788/2020\|EPI_ISL_465685\|20 | ···················· | ···················· | ···················· | ···················· | ···················· | ···················· |
| D | 4713 | hCoV-19/Canada/Qc-L00240796/2020\|EPI_ISL_465686\|20 | ···················· | ···················· | ···················· | ···················· | ···················· | ···················· |
| D | 4714 | hCoV-19/Canada/Qc-L00240804/2020\|EPI_ISL_465687\|20 | ···················· | ···················· | ···················· | ···················· | ···················· | ···················· |
| D | 4715 | hCoV-19/Canada/Qc-L00240846/2020\|EPI_ISL_465688\|20 | ···················· | ···················· | ···················· | ···················· | ···················· | ···················· |
| D | 4716 | hCoV-19/Canada/Qc-L00240851/2020\|EPI_ISL_465689\|20 | ···················· | ···················· | ···················· | ···················· | ···················· | ···················· |
| D | 4717 | hCoV-19/Canada/Qc-L00240866/2020\|EPI_ISL_465690\|20 | ···················· | ···················· | ···················· | ···················· | ···················· | ···················· |
| D | 4718 | hCoV-19/Canada/Qc-L00240891/2020\|EPI_ISL_465691\|20 | ···················· | ···················· | ···················· | ···················· | ···················· | ···················· |
| D | 4719 | hCoV-19/Canada/Qc-L00240893/2020\|EPI_ISL_465692\|20 | ···················· | ···················· | ···················· | ···················· | ···················· | ···················· |
| D | 4720 | hCoV-19/Canada/Qc-L00240947/2020\|EPI_ISL_465693\|20 | ···················· | ···················· | ···················· | ···················· | ···················· | ···················· |
| D | 4721 | hCoV-19/Canada/Qc-L00241007/2020\|EPI_ISL_465694\|20 | ···················· | ···················· | ···················· | ···················· | ···················· | ···················· |
| D | 4722 | hCoV-19/Canada/Qc-L00241074/2020\|EPI_ISL_465695\|20 | ···················· | ···················· | ···················· | ···················· | ···················· | ···················· |
| D | 4723 | hCoV-19/Canada/Qc-L00241102/2020\|EPI_ISL_465696\|20 | ···················· | ···················· | ···················· | ···················· | ···················· | ···················· |
| D | 4724 | hCoV-19/Canada/Qc-L00241112/2020\|EPI_ISL_465697\|20 | ···················· | ···················· | ···················· | ···················· | ···················· | ···················· |
| D | 4725 | hCoV-19/Canada/Qc-L00241115/2020\|EPI_ISL_465698\|20 | ···················· | ···················· | ···················· | ···················· | ···················· | ···················· |
| D | 4726 | hCoV-19/Canada/Qc-L00241158/2020\|EPI_ISL_465699\|20 | ···················· | ···················· | ···················· | ···················· | ···················· | ···················· |
| D | 4727 | hCoV-19/England/201580189/2020\|EPI_ISL_466359\|2020 | ···················· | ···················· | ···················· | ···················· | ···················· | ···················· |
| D | 4728 | hCoV-19/England/201580190/2020\|EPI_ISL_466360\|2020 | ···················· | ···················· | ···················· | ···················· | ···················· | ···················· |
| D | 4729 | hCoV-19/England/201580290/2020\|EPI_ISL_466367\|2020 | ···················· | ···················· | ···················· | ···················· | ···················· | ···················· |
| D | 4730 | hCoV-19/Spain/CM-IBV-003760/2020\|EPI_ISL_474803\|20 | ···················· | ···················· | ···················· | ···················· | ···················· | ···················· |
| D | 4737 | hCoV-19/India/GJ-GBRC203a/2020\|EPI_ISL_475026\|2020 | ···················· | ···················· | ···················· | ···················· | ···················· | ···················· |
| D | 4738 | hCoV-19/India/GJ-GBRC203b/2020\|EPI_ISL_475027\|2020 | ···················· | ·A·················· | ···················· | ···················· | ···················· | ··A················· |
| D | 4739 | hCoV-19/India/GJ-GBRC204a/2020\|EPI_ISL_475028\|2020 | ···················· | ·A·················· | ···················· | ···················· | ···················· | ··A················· |
| D | 4740 | hCoV-19/India/GJ-GBRC204b/2020\|EPI_ISL_475029\|2020 | ···T················ | ·A·················· | ···················· | ···················· | ···················· | ··A················· |
| D | 4741 | hCoV-19/India/GJ-GBRC205a/2020\|EPI_ISL_475030\|2020 | ···················· | ·A·················· | ···················· | ···················· | ···················· | ··A················· |
| D | 4742 | hCoV-19/India/GJ-GBRC205b/2020\|EPI_ISL_475031\|2020 | ···················· | ·A·················· | ···················· | ···················· | ···················· | ··A················· |
| D | 4743 | hCoV-19/India/GJ-GBRC206/2020\|EPI_ISL_475032\|2020- | ···················· | ···················· | ···················· | ···················· | ···················· | ···················· |
| D | 4744 | hCoV-19/India/GJ-GBRC207a/2020\|EPI_ISL_475033\|2020 | ···················· | ·A·················· | ···················· | ···················· | ···················· | ··A················· |
| D | 4745 | hCoV-19/India/GJ-GBRC207b/2020\|EPI_ISL_475034\|2020 | ···················· | ···················· | ···················· | ···················· | ···················· | ··A················· |
| D | 4746 | hCoV-19/India/GJ-GBRC208a/2020\|EPI_ISL_475035\|2020 | ···················· | ···················· | ···················· | ···················· | ···················· | ···················· |
| D | 4747 | hCoV-19/India/GJ-GBRC208b/2020\|EPI_ISL_475036\|2020 | ···················· | ·A·················· | ···················· | ···················· | ···················· | ··A················· |
| D | 4748 | hCoV-19/India/GJ-GBRC209a/2020\|EPI_ISL_475037\|2020 | ···················· | ···················· | ···················· | ···················· | ···················· | ···················· |
| D | 4749 | hCoV-19/India/GJ-GBRC209b/2020\|EPI_ISL_475038\|2020 | ···················· | ·A·················· | ···················· | ···················· | ···················· | ··A················· |
| D | 4750 | hCoV-19/India/GJ-GBRC210a/2020\|EPI_ISL_475039\|2020 | ···················· | ·A·················· | ···················· | ···················· | ···················· | ··A················· |
| D | 4751 | hCoV-19/India/GJ-GBRC210b/2020\|EPI_ISL_475040\|2020 | ···················· | ···················· | ···················· | ···················· | ···················· | ···················· |
| D | 4752 | hCoV-19/India/GJ-GBRC213a/2020\|EPI_ISL_475043\|2020 | ···················· | ···················· | ···················· | ···················· | ···················· | ··A················· |
| D | 4753 | hCoV-19/India/GJ-GBRC213b/2020\|EPI_ISL_475044\|2020 | ···················· | ···················· | ···················· | ···················· | ···················· | ···················· |
| D | 4754 | hCoV-19/India/GJ-GBRC214a/2020\|EPI_ISL_475045\|2020 | ···················· | ···················· | ···················· | ···················· | ······T············· | ···················· |
| D | 4755 | hCoV-19/India/GJ-GBRC214b/2020\|EPI_ISL_475046\|2020 | ···················· | ·A·················· | ···················· | ···················· | ···················· | ··A················· |
| D | 4756 | hCoV-19/India/GJ-GBRC194a/2020\|EPI_ISL_475047\|2020 | ···················· | ·A·················· | ···················· | ···················· | ···················· | ··A················· |
| D | 4757 | hCoV-19/India/GJ-GBRC194b/2020\|EPI_ISL_475048\|2020 | ···················· | ·A·················· | ···················· | ···················· | ···················· | ··A················· |
| D | 4758 | hCoV-19/India/GJ-GBRC215/2020\|EPI_ISL_475049\|2020- | ···················· | ···················· | ···················· | ···················· | ···················· | ··A················· |
| D | 4759 | hCoV-19/India/GJ-GBRC216a/2020\|EPI_ISL_475050\|2020 | ···················· | ·A·················· | ···················· | ···················· | ···················· | ··A················· |
| D | 4760 | hCoV-19/India/GJ-GBRC216b/2020\|EPI_ISL_475051\|2020 | ···················· | ·A·················· | ···················· | ···················· | ···················· | ··A················· |
| D | 4761 | hCoV-19/India/GJ-GBRC217a/2020\|EPI_ISL_475052\|2020 | ···················· | ·A·················· | ···················· | ···················· | ···················· | ··A················· |
| D | 4762 | hCoV-19/India/GJ-GBRC217b/2020\|EPI_ISL_475053\|2020 | ···················· | ···················· | ···················· | ···················· | ···················· | ··A················· |
| D | 4763 | hCoV-19/India/GJ-GBRC218a/2020\|EPI_ISL_475054\|2020 | ···················· | ·A·················· | ···················· | ···················· | ···················· | ··A················· |
| D | 4764 | hCoV-19/India/GJ-GBRC218b/2020\|EPI_ISL_475055\|2020 | ···················· | ·A·················· | ···················· | ···················· | ···················· | ··A················· |
| D | 4765 | hCoV-19/India/GJ-GBRC219a/2020\|EPI_ISL_475056\|2020 | ···················· | ·A·················· | ···················· | ···················· | ···················· | ··A················· |
| D | 4766 | hCoV-19/India/GJ-GBRC219b/2020\|EPI_ISL_475057\|2020 | ···················· | ·A·················· | ···················· | ···················· | ···················· | ··A················· |
| D | 4767 | hCoV-19/India/GJ-GBRC220/2020\|EPI_ISL_475058\|2020- | ···················· | ···················· | ···················· | ···················· | ···················· | ···················· |
| D | 4768 | hCoV-19/India/GJ-GBRC221/2020\|EPI_ISL_475059\|2020- | ···················· | ···················· | ···················· | ···················· | ···················· | ···················· |
| D | 4769 | hCoV-19/Belgium/UGent-100/2020\|EPI_ISL_475060\|2020 | ···················· | ···················· | ···················· | ···················· | ···················· | ···················· |
| D | 4771 | hCoV-19/Brazil/SP-L22-CD474/2020\|EPI_ISL_476373\|20 | ···················· | ···················· | ···················· | ···················· | ···················· | ···················· |
| D | 4772 | hCoV-19/Belgium/rega-0511429/2020\|EPI_ISL_477006\|2 | ···················· | ···················· | ···················· | ···················· | ···················· | ···················· |
| D | 4773 | hCoV-19/Belgium/rega-0513430/2020\|EPI_ISL_477007\|2 | ···················· | ···················· | ···················· | ···················· | ···················· | ···················· |
| D | 4774 | hCoV-19/Hungary/UD-10168/2020\|EPI_ISL_477008\|2020- | ···················· | ···················· | ···················· | ···················· | ···················· | ···················· |
| D | 4775 | hCoV-19/Hungary/UD-10333/2020\|EPI_ISL_477009\|2020- | ···················· | ···················· | ···················· | ···················· | ···················· | ···················· |
| D | 4776 | hCoV-19/Hungary/UD-6802/2020\|EPI_ISL_477010\|2020-0 | ···················· | ···················· | ···················· | ···················· | ···················· | ···················· |
| D | 4777 | hCoV-19/Hungary/UD-7360/2020\|EPI_ISL_477011\|2020-0 | ···················· | ···················· | ···················· | ···················· | ···················· | ···················· |
| D | 4778 | hCoV-19/Hungary/UD-7364/2020\|EPI_ISL_477012\|2020-0 | ···················· | ···················· | ···················· | ···················· | ···················· | ···················· |
| D | 4779 | hCoV-19/Hungary/UD-9953/2020\|EPI_ISL_477013\|2020-0 | ···················· | ···················· | ···················· | ···················· | ···················· | ···················· |
| D | 4780 | hCoV-19/Ecuador/USFQ-004/2020\|EPI_ISL_477014\|2020- | ···················· | ···················· | ···················· | ···················· | ···················· | ···················· |
| D | 4781 | hCoV-19/Ecuador/USFQ-096/2020\|EPI_ISL_477015\|2020- | ···················· | ···················· | ···················· | ···················· | ···················· | ···················· |

| | | | | | | | | |
|---|---|---|---|---|---|---|---|---|
| D | 4782 | hCoV-19/Ecuador/USFQ-097/2020\|EPI_ISL_477016\|2020- | ···················· | ···················· | ···················· | ···················· | ···················· | ···················· |
| D | 4783 | hCoV-19/Canada/BC_19909443/2020\|EPI_ISL_477020\|202 | ···················· | ···················· | ···················· | ···················· | ···················· | ···················· |
| D | 4784 | hCoV-19/Canada/BC_19909482/2020\|EPI_ISL_477021\|202 | ···················· | ···················· | ···················· | ···················· | ···················· | ···················· |
| D | 4785 | hCoV-19/Canada/BC_20367856/2020\|EPI_ISL_477022\|202 | ···················· | ···················· | ···················· | ···················· | ···················· | ···················· |
| D | 4786 | hCoV-19/Canada/BC_21355315/2020\|EPI_ISL_477023\|202 | ···················· | ···················· | ···················· | ···················· | ···················· | ···················· |
| D | 4787 | hCoV-19/Canada/BC_21358214/2020\|EPI_ISL_477024\|202 | ···················· | ···················· | ···················· | ···················· | ···················· | ···················· |
| D | 4788 | hCoV-19/Canada/BC_21451231/2020\|EPI_ISL_477029\|202 | ···················· | ···················· | ···················· | ···················· | ···················· | ···················· |
| D | 4789 | hCoV-19/Canada/BC_22422949/2020\|EPI_ISL_477031\|202 | ···················· | ···················· | ···················· | ···················· | ···················· | ···················· |
| D | 4790 | hCoV-19/Canada/BC_22502164/2020\|EPI_ISL_477032\|202 | ···················· | ···················· | ···················· | ···················· | ···················· | ···················· |
| D | 4791 | hCoV-19/Canada/BC_22510360/2020\|EPI_ISL_477033\|202 | ···················· | ···················· | ···················· | ···················· | ···················· | ···················· |
| D | 4792 | hCoV-19/Canada/BC_22611474/2020\|EPI_ISL_477034\|202 | ···················· | ···················· | ···················· | ···················· | ···················· | ···················· |
| D | 4793 | hCoV-19/Canada/BC_23246810/2020\|EPI_ISL_477035\|202 | ···················· | ···················· | ···················· | ···················· | ···················· | ···················· |
| D | 4794 | hCoV-19/Canada/BC_23252816/2020\|EPI_ISL_477036\|202 | ···················· | ···················· | ···················· | ···················· | ···················· | ···················· |
| D | 4795 | hCoV-19/Canada/BC_23279341/2020\|EPI_ISL_477037\|202 | ···················· | ···················· | ···················· | ···················· | ···················· | ···················· |
| D | 4796 | hCoV-19/Canada/BC_26060645/2020\|EPI_ISL_477038\|202 | ···················· | ···················· | ···················· | ···················· | ···················· | ···················· |
| D | 4797 | hCoV-19/Canada/BC_00740314/2020\|EPI_ISL_477039\|202 | ···················· | ···················· | ···················· | ···················· | ···················· | ···················· |
| D | 4798 | hCoV-19/Canada/BC_01775252/2020\|EPI_ISL_477040\|202 | ···················· | ···················· | ···················· | ···················· | ···················· | ···················· |
| D | 4799 | hCoV-19/Canada/BC_02281789/2020\|EPI_ISL_477041\|202 | ···················· | ···················· | ···················· | ···················· | ···················· | ···················· |
| D | 4800 | hCoV-19/Canada/BC_04547356/2020\|EPI_ISL_477045\|202 | ···················· | ···················· | ···················· | ···················· | ···················· | ···················· |
| D | 4801 | hCoV-19/Canada/BC_04579295/2020\|EPI_ISL_477046\|202 | ···················· | ···················· | ···················· | ···················· | ···················· | ···················· |
| D | 4802 | hCoV-19/Canada/BC_04583818/2020\|EPI_ISL_477047\|202 | ···················· | ···················· | ···················· | ···················· | ···················· | ···················· |
| D | 4803 | hCoV-19/Canada/BC_04677077/2020\|EPI_ISL_477048\|202 | ···················· | ···················· | ···················· | ···················· | ···················· | ···················· |
| D | 4804 | hCoV-19/Canada/BC_04684452/2020\|EPI_ISL_477049\|202 | ···················· | ···················· | ···················· | ···················· | ···················· | ···················· |
| D | 4805 | hCoV-19/Canada/BC_10420892/2020\|EPI_ISL_477080\|202 | ···················· | ···················· | ···················· | ···················· | ···················· | ···················· |
| D | 4806 | hCoV-19/Canada/BC_13929277/2020\|EPI_ISL_477081\|202 | ···················· | ···················· | ···················· | ···················· | ···················· | ···················· |
| D | 4807 | hCoV-19/Canada/BC_16505592/2020\|EPI_ISL_477085\|202 | ···················· | ···················· | ···················· | ···················· | ···················· | ···················· |
| D | 4808 | hCoV-19/Canada/BC_16510082/2020\|EPI_ISL_477086\|202 | ···················· | ···················· | ···················· | ···················· | ···················· | ···················· |
| D | 4809 | hCoV-19/Canada/BC_02018462/2020\|EPI_ISL_477087\|202 | ···················· | ···················· | ···················· | ···················· | ···················· | ···················· |
| D | 4810 | hCoV-19/Canada/BC_02038211/2020\|EPI_ISL_477088\|202 | ···················· | ···················· | ···················· | ···················· | ···················· | ···················· |
| D | 4811 | hCoV-19/USA/WA-UW-1613/2020\|EPI_ISL_477689\|2020-03 | ···················· | ···················· | ···················· | ···················· | ···················· | ···················· |
| D | 4812 | hCoV-19/USA/WA-UW-1617/2020\|EPI_ISL_477690\|2020-03 | ···················· | ···················· | ···················· | ···················· | ···················· | ···················· |
| D | 4813 | hCoV-19/USA/WA-UW-1624/2020\|EPI_ISL_477691\|2020-03 | ···················· | ···················· | ···················· | ···················· | ···················· | ···················· |
| D | 4814 | hCoV-19/USA/WA-UW-1630/2020\|EPI_ISL_477692\|2020-03 | ···················· | ···················· | ···················· | ···················· | ···················· | ···················· |
| D | 4815 | hCoV-19/USA/WA-UW-1631/2020\|EPI_ISL_477693\|2020-03 | ···················· | ···················· | ···················· | ···················· | ···················· | ···················· |
| D | 4816 | hCoV-19/USA/WA-UW-1632/2020\|EPI_ISL_477694\|2020-03 | ···················· | ···················· | ···················· | ···················· | ···················· | ···················· |
| D | 4817 | hCoV-19/USA/WA-UW-2032/2020\|EPI_ISL_477695\|2020-03 | ···················· | ···················· | ···················· | ···················· | ···················· | ···················· |
| D | 4818 | hCoV-19/USA/WA-UW-2036/2020\|EPI_ISL_477696\|2020-03 | ···················· | ···················· | ···················· | ···················· | ···················· | ···················· |
| D | 4819 | hCoV-19/USA/WA-UW-2039/2020\|EPI_ISL_477697\|2020-03 | ···················· | ···················· | ···················· | ···················· | ···················· | ···················· |
| D | 4820 | hCoV-19/USA/WA-UW-2045/2020\|EPI_ISL_477698\|2020-03 | ···················· | ···················· | ···················· | ···················· | ···················· | ···················· |
| D | 4821 | hCoV-19/USA/WA-UW-2048/2020\|EPI_ISL_477699\|2020-03 | ···················· | ···················· | ···················· | ···················· | ···················· | ···················· |
| D | 4822 | hCoV-19/England/LCST-24D6C8D/2020\|EPI_ISL_478338\|2 | ···················· | ···················· | ···················· | ···················· | ···················· | ···················· |
| D | 4823 | hCoV-19/England/LCST-24D6E87/2020\|EPI_ISL_478343\|2 | ···················· | ···················· | ···················· | ···················· | ···················· | ···················· |
| D | 4824 | hCoV-19/England/LCST-24D6F84/2020\|EPI_ISL_478345\|2 | ···················· | ···················· | ···················· | ···················· | ···················· | ···················· |
| D | 4825 | hCoV-19/England/LCST-24D6FB1/2020\|EPI_ISL_478347\|2 | ···················· | ···················· | ···················· | ···················· | ···················· | ···················· |
| D | 4826 | hCoV-19/England/LCST-24D6FDF/2020\|EPI_ISL_478349\|2 | ···················· | ···················· | ···················· | ···················· | ···················· | ···················· |
| D | 4827 | hCoV-19/England/LCST-24D6FEE/2020\|EPI_ISL_478350\|2 | ···················· | ···················· | ···················· | ···················· | ···················· | ···················· |
| D | 4828 | hCoV-19/England/LCST-24D6FFD/2020\|EPI_ISL_478351\|2 | ···················· | ···················· | ···················· | ···················· | ···················· | ···················· |
| D | 4829 | hCoV-19/England/LCST-24D7017/2020\|EPI_ISL_478353\|2 | ···················· | ···················· | ···················· | ···················· | ···················· | ···················· |
| D | 4830 | hCoV-19/England/LCST-24D7035/2020\|EPI_ISL_478355\|2 | ···················· | ···················· | ···················· | ···················· | ···················· | ···················· |
| D | 4831 | hCoV-19/England/LCST-24D7080/2020\|EPI_ISL_478359\|2 | ···················· | ···················· | ···················· | ···················· | ···················· | ···················· |
| D | 4832 | hCoV-19/England/LCST-24D709F/2020\|EPI_ISL_478360\|2 | ···················· | ···················· | ···················· | ···················· | ···················· | ···················· |
| D | 4833 | hCoV-19/England/LCST-24D70AE/2020\|EPI_ISL_478361\|2 | ···················· | ···················· | ···················· | ···················· | ···················· | ···················· |
| D | 4834 | hCoV-19/England/LCST-24D70BD/2020\|EPI_ISL_478362\|2 | ···················· | ···················· | ···················· | ···················· | ···················· | ···················· |
| D | 4835 | hCoV-19/England/LCST-24D718D/2020\|EPI_ISL_478370\|2 | ···················· | ···················· | ···················· | ···················· | ···················· | ···················· |
| D | 4836 | hCoV-19/England/LCST-24D71AB/2020\|EPI_ISL_478371\|2 | ···················· | ···················· | ···················· | ···················· | ···················· | ···················· |
| D | 4837 | hCoV-19/USA/NY-NYU-VC-014/2020\|EPI_ISL_479668\|2020 | ···················· | ···················· | ···················· | ···················· | ···················· | ···················· |
| D | 4838 | hCoV-19/USA/NY-NYU-VC-017/2020\|EPI_ISL_479671\|2020 | ···················· | ···················· | ···················· | ···················· | ···················· | ···················· |
| D | 4839 | hCoV-19/USA/NY-NYU-VC-018/2020\|EPI_ISL_479672\|2020 | ···················· | ···················· | ···················· | ···················· | ···················· | ···················· |
| D | 4840 | hCoV-19/USA/NY-NYU-VC-022/2020\|EPI_ISL_479674\|2020 | ···················· | ···················· | ···················· | ···················· | ···················· | ···················· |
| D | 4841 | hCoV-19/Japan/TKYE65734/2020\|EPI_ISL_479676\|2020-0 | ···················· | ···················· | ···················· | ···················· | ···················· | ···················· |
| D | 4842 | hCoV-19/Japan/TKYE610419/2020\|EPI_ISL_479677\|2020- | ···················· | ···················· | ···················· | ···················· | ···················· | ···················· |
| D | 4843 | hCoV-19/Japan/TKYE610497/2020\|EPI_ISL_479678\|2020- | ···················· | ···················· | ···················· | ···················· | ···················· | ···················· |
| D | 4844 | hCoV-19/Japan/TKYE610504/2020\|EPI_ISL_479679\|2020- | ···················· | ···················· | ···················· | ···················· | ···················· | ···················· |
| D | 4845 | hCoV-19/Japan/TKYE610510/2020\|EPI_ISL_479680\|2020- | ···················· | ···················· | ···················· | ···················· | ···················· | ···················· |
| D | 4846 | hCoV-19/Japan/TKYE610670/2020\|EPI_ISL_479681\|2020- | ···················· | ···················· | ···················· | ···················· | ···················· | ···················· |
| D | 4847 | hCoV-19/Japan/TKYE610850/2020\|EPI_ISL_479682\|2020- | ···················· | ···················· | ···················· | ···················· | ···················· | ···················· |
| D | 4848 | hCoV-19/Japan/TKYVE11964/2020\|EPI_ISL_479683\|2020- | ···················· | ···················· | ···················· | ···················· | ···················· | ···················· |
| D | 4849 | hCoV-19/Japan/TKYVE12492/2020\|EPI_ISL_479684\|2020- | ···················· | ···················· | ···················· | ···················· | ···················· | ···················· |
| D | 4850 | hCoV-19/Japan/TKYE613173/2020\|EPI_ISL_479685\|2020- | ···················· | ···················· | ···················· | ···················· | ···················· | ···················· |
| D | 4851 | hCoV-19/Egypt/CUNCI-HGC3I02/2020\|EPI_ISL_479686\|20 | ···················· | ·····R·············· | ···················· | ···················· | ···················· | ···················· |
| D | 4852 | hCoV-19/Egypt/CUNCI-HGC3I03/2020\|EPI_ISL_479687\|20 | ···················· | ·····A·············· | ···················· | ···················· | ···················· | ···················· |
| D | 4853 | hCoV-19/Egypt/CUNCI-HGC3I06/2020\|EPI_ISL_479688\|20 | ···················· | ·····A·············· | ···················· | ···················· | ···················· | ···················· |
| D | 4854 | hCoV-19/Egypt/CUNCI-HGC3I07/2020\|EPI_ISL_479689\|20 | ···················· | ·····A·············· | ···················· | ···················· | ···················· | ···················· |
| D | 4855 | hCoV-19/Egypt/CUNCI-HGC3I05/2020\|EPI_ISL_479690\|20 | ···················· | ···················· | ···················· | ···················· | ···················· | ···················· |
| D | 4856 | hCoV-19/Egypt/CUNCI-HGC3I025/2020\|EPI_ISL_479691\|2 | ···················· | ···················· | ···················· | ···················· | ···················· | ···················· |
| D | 4857 | hCoV-19/Egypt/CUNCI-HGC3I09/2020\|EPI_ISL_479692\|20 | ···················· | ···················· | ···················· | ···················· | ···················· | ···················· |
| D | 4858 | hCoV-19/Egypt/CUNCI-HGC3I012/2020\|EPI_ISL_479693\|2 | ···················· | ···················· | ···················· | ···················· | ···················· | ···················· |
| D | 4859 | hCoV-19/Egypt/CUNCI-HGC3I013/2020\|EPI_ISL_479694\|2 | ···················· | ···················· | ···················· | ···················· | ···················· | ···················· |

| | | | | | | | | |
|---|---|---|---|---|---|---|---|---|
| D | 4860 hCoV-19/Egypt/CUNCI-HGC3I023/2020\|EPI_ISL_479695\|2 | ···················· | ······R············· | ···················· | ···················· | ···················· | ···················· | |
| D | 4861 hCoV-19/Egypt/CUNCI-HGC3I014/2020\|EPI_ISL_479696\|2 | ···················· | ···················· | ···················· | ···················· | ···················· | ···················· | |
| D | 4862 hCoV-19/Egypt/CUNCI-HGC3I011/2020\|EPI_ISL_479697\|2 | ···················· | ···················· | ···················· | ···················· | ···················· | ···················· | |
| D | 4863 hCoV-19/Egypt/CUNCI-HGC3I08/2020\|EPI_ISL_479698\|20 | ···················· | ······A············· | ···················· | ···················· | ···················· | ···················· | |
| D | 4864 hCoV-19/Egypt/CUNCI-HGC4I003/2020\|EPI_ISL_479699\|2 | ···················· | ···················· | ···················· | ···················· | ···················· | ···················· | |
| D | 4865 hCoV-19/Taiwan/NTU29/2020\|EPI_ISL_463007\|2020-04-1 | ···················· | ···················· | ···················· | ···················· | ···················· | ···················· | |
| D | 4866 hCoV-19/Germany/NW-IMVUM_FI1103201/2020\|EPI_ISL_46 | ···················· | ···················· | ···················· | ···················· | ···················· | ···················· | |
| D | 4867 hCoV-19/India/OR-ILSCV16509/2020\|EPI_ISL_463049\|20 | ···················· | ···················· | ···················· | ···················· | ···················· | ···················· | |
| D | 4868 hCoV-19/India/OR-ILSCV16512/2020\|EPI_ISL_463051\|20 | ···················· | ···················· | ···················· | ···················· | ···················· | ···················· | |
| D | 4869 hCoV-19/India/OR-ILSCV16514/2020\|EPI_ISL_463052\|20 | ···················· | ···················· | ···················· | ···················· | ···················· | ···················· | |
| D | 4870 hCoV-19/India/OR-ILSCV16809/2020\|EPI_ISL_463054\|20 | ···················· | ···················· | ···················· | ···················· | ···················· | ···················· | |
| D | 4871 hCoV-19/India/OR-ILSCV17568/2020\|EPI_ISL_463056\|20 | ···················· | ···················· | ···················· | ···················· | ···················· | ···················· | |
| D | 4872 hCoV-19/India/OR-ILSCV17741/2020\|EPI_ISL_463058\|20 | ···················· | ···················· | ···················· | ···················· | ···················· | ···················· | |
| D | 4873 hCoV-19/India/OR-ILSCV18525/2020\|EPI_ISL_463060\|20 | ···················· | ·A·················· | ···················· | ···················· | ···················· | ···A················ | |
| D | 4874 hCoV-19/India/OR-ILSCV18534/2020\|EPI_ISL_463061\|20 | ···················· | ·A·················· | ···················· | ···················· | ···················· | ···A················ | |
| D | 4876 hCoV-19/India/OR-ILSCV19048/2020\|EPI_ISL_463064\|20 | ···················· | ···················· | ···················· | ···················· | ···················· | ···················· | |
| D | 4877 hCoV-19/India/OR-ILSCV19192/2020\|EPI_ISL_463067\|20 | ···················· | ···················· | ···················· | ···················· | ···················· | ···················· | |
| D | 4878 hCoV-19/India/OR-ILSCV20165/2020\|EPI_ISL_463073\|20 | ···················· | ···················· | ···················· | ···················· | ···················· | ···················· | |
| D | 4879 hCoV-19/India/OR-ILSCV20166/2020\|EPI_ISL_463074\|20 | ···················· | ···················· | ···················· | ···················· | ···················· | ···················· | |
| D | 4880 hCoV-19/India/OR-ILSCV20408/2020\|EPI_ISL_463076\|20 | ···················· | ···················· | ···················· | ···················· | ···················· | ···················· | |
| D | 4881 hCoV-19/India/OR-ILSCV20557/2020\|EPI_ISL_463078\|20 | ···················· | ···················· | ···················· | ···················· | ···················· | ···················· | |
| D | 4882 hCoV-19/India/OR-ILSCV20600/2020\|EPI_ISL_463081\|20 | ···················· | ···················· | ···················· | ···················· | ···················· | ···················· | |
| D | 4883 hCoV-19/India/OR-ILSCV21721/2020\|EPI_ISL_463085\|20 | ···················· | ···················· | ···················· | ···················· | ···················· | ···················· | |
| D | 4884 hCoV-19/India/OR-ILSCV24694/2020\|EPI_ISL_463087\|20 | ···················· | ···················· | ···················· | ···················· | ···················· | ···················· | |
| D | 4885 hCoV-19/India/OR-ILSCV24917/2020\|EPI_ISL_463088\|20 | ···················· | ···················· | ···················· | ···················· | ···················· | ···················· | |
| D | 4886 hCoV-19/India/OR-ILSCV24918/2020\|EPI_ISL_463089\|20 | ···················· | ···················· | ···················· | ···················· | ···················· | ···················· | |
| D | 4887 hCoV-19/England/201040152/2020\|EPI_ISL_464334\|2020 | ···················· | ···················· | ···················· | ···················· | ···················· | ···················· | |
| D | 4888 hCoV-19/England/201040154/2020\|EPI_ISL_464335\|2020 | ···················· | ···················· | ···················· | ···················· | ···················· | ···················· | |
| D | 4889 hCoV-19/England/201040375/2020\|EPI_ISL_464339\|2020 | ···················· | ······C············· | ···················· | ···················· | ···················· | ···················· | |
| D | 4891 hCoV-19/England/201060046/2020\|EPI_ISL_464380\|2020 | ···················· | ···················· | ···················· | ···················· | ···················· | ···················· | |
| D | 4892 hCoV-19/England/201060050/2020\|EPI_ISL_464382\|2020 | ···················· | ···················· | ···················· | ···················· | ··············N····N | ·····Y·············· | 1 |
| D | 4893 hCoV-19/England/201060052/2020\|EPI_ISL_464383\|2020 | ···················· | ···················· | ···················· | ···················· | ···················· | ···················· | |
| D | 4894 hCoV-19/England/201060059/2020\|EPI_ISL_464388\|2020 | ···················· | ···················· | ···················· | ···················· | ··N················· | ··N······N······N··· | 1 |
| D | 4895 hCoV-19/England/201060061/2020\|EPI_ISL_464389\|2020 | ···················· | ···················· | ···················· | ···················· | ···················· | ···················· | |
| D | 4899 hCoV-19/England/201060142/2020\|EPI_ISL_464395\|2020 | ···················· | ···················· | ···················· | ···················· | ···················· | ···················· | |
| D | 4901 hCoV-19/England/201060144/2020\|EPI_ISL_464397\|2020 | ···················· | ···················· | ···················· | ···················· | ···················· | ···················· | |
| D | 4902 hCoV-19/England/201060160/2020\|EPI_ISL_464399\|2020 | ···················· | ···················· | ···················· | ···················· | ···················· | ···················· | |
| D | 4904 hCoV-19/England/20192083901/2020\|EPI_ISL_465601\|20 | ···················· | ···················· | ···················· | ···················· | ···················· | ··N················· | |
| D | 4905 hCoV-19/England/20192085601/2020\|EPI_ISL_465602\|20 | ···················· | ···················· | ···················· | ···················· | ···················· | ···················· | |
| D | 4907 hCoV-19/England/20192179501/2020\|EPI_ISL_465605\|20 | ···················· | ···················· | ···················· | ···················· | ···················· | ···················· | |
| D | 4910 hCoV-19/England/20194041601/2020\|EPI_ISL_465610\|20 | ···················· | ···················· | ···················· | ···················· | ···················· | ···················· | |
| D | 4911 hCoV-19/England/20194099704/2020\|EPI_ISL_465611\|20 | ···················· | ···················· | ···················· | ···················· | ···················· | ···················· | |
| D | 4912 hCoV-19/England/20194102101/2020\|EPI_ISL_465612\|20 | ···················· | ···················· | ···················· | ···················· | ···················· | ···················· | |
| D | 4915 hCoV-19/England/20194106101/2020\|EPI_ISL_465615\|20 | ···················· | ···················· | ···················· | ···················· | ···················· | ···················· | |
| D | 4916 hCoV-19/England/20194106301/2020\|EPI_ISL_465616\|20 | ···················· | ···················· | ···················· | ···················· | ···················· | ···················· | |
| D | 4917 hCoV-19/England/20194106601/2020\|EPI_ISL_465617\|20 | ···················· | ···················· | ···················· | ···················· | ···················· | ··N················· | |
| D | 4919 hCoV-19/England/20194148101/2020\|EPI_ISL_465620\|20 | ···················· | ···················· | ···················· | ···················· | ···················· | ···················· | |
| D | 4922 hCoV-19/England/20206140304/2020\|EPI_ISL_465668\|20 | ···················· | ···················· | ···················· | ···················· | ···················· | ···················· | |
| D | 4923 hCoV-19/Germany/BY-MVP-0288/2020\|EPI_ISL_466916\|20 | ···················· | ···················· | ···················· | ···················· | ···················· | ···················· | |
| D | 4924 hCoV-19/Germany/BY-MVP-0284/2020\|EPI_ISL_466917\|20 | ···················· | ···················· | ···················· | ···················· | ···················· | ···················· | |
| D | 4925 hCoV-19/Germany/BY-MVP-0272/2020\|EPI_ISL_466918\|20 | ···················· | ···················· | ···················· | ···················· | ···················· | ···················· | |
| D | 4926 hCoV-19/Germany/BY-MVP-0276/2020\|EPI_ISL_466919\|20 | ···················· | ···················· | ···················· | ···················· | ···················· | ···················· | |
| D | 4927 hCoV-19/Germany/BY-MVP-0277/2020\|EPI_ISL_466920\|20 | ···················· | ···················· | ···················· | ···················· | ···················· | ···················· | |
| D | 4928 hCoV-19/Germany/BY-MVP-0287/2020\|EPI_ISL_466921\|20 | ···················· | ···················· | ···················· | ···················· | ···················· | ···················· | |
| D | 4929 hCoV-19/Germany/BY-MVP-0289/2020\|EPI_ISL_466922\|20 | ···················· | ···················· | ···················· | ···················· | ···················· | ···················· | |
| D | 4930 hCoV-19/Germany/BY-MVP-0291/2020\|EPI_ISL_466923\|20 | ···················· | ···················· | ···················· | ···················· | ···················· | ···················· | |
| D | 4931 hCoV-19/Germany/BY-MVP-0294/2020\|EPI_ISL_466924\|20 | ···················· | ···················· | ···················· | ···················· | ···················· | ···················· | |
| D | 4932 hCoV-19/Germany/BY-MVP-0296/2020\|EPI_ISL_466925\|20 | ···················· | ···················· | ···················· | ···················· | ···················· | ···················· | |
| D | 4933 hCoV-19/Switzerland/BS-ETHZ-100002/2020\|EPI_ISL_46 | ···················· | ···················· | ···················· | ···················· | ·······M············ | ···················· | |
| D | 4938 hCoV-19/Switzerland/GR-ETHZ-100007/2020\|EPI_ISL_46 | ···················· | ···················· | ···················· | ···················· | ···················· | ···················· | |
| D | 4941 hCoV-19/Switzerland/TI-ETHZ-100011/2020\|EPI_ISL_46 | ···················· | ···················· | ···················· | ···················· | ···················· | ···················· | |
| D | 4942 hCoV-19/Switzerland/TI-ETHZ-100012/2020\|EPI_ISL_46 | ···················· | ···················· | ···················· | ···················· | ···················· | ··K················· | |
| D | 4945 hCoV-19/Switzerland/BS-ETHZ-100016/2020\|EPI_ISL_46 | ·········K·········· | ···················· | ···················· | ···················· | ·······S············ | ···················· | |
| D | 4948 hCoV-19/Switzerland/SO-ETHZ-100019/2020\|EPI_ISL_46 | ···················· | ···················· | ···················· | ···················· | ···················· | ···················· | |
| D | 4953 hCoV-19/Switzerland/BS-ETHZ-100025/2020\|EPI_ISL_46 | ···················· | ···················· | ···················· | ···················· | ············N··N···· | ···················· | |
| D | 4954 hCoV-19/Switzerland/GR-ETHZ-100026/2020\|EPI_ISL_46 | ···················· | ···················· | ···················· | ···················· | ···················· | ···················· | |
| D | 4955 hCoV-19/Switzerland/BS-ETHZ-100027/2020\|EPI_ISL_46 | ···················· | ···················· | ···················· | ···················· | ·············N······ | ···················· | |
| D | 4959 hCoV-19/USA/CT-Yale-270/2020\|EPI_ISL_469194\|2020-0 | ···················· | ···················· | ···················· | ···················· | ···················· | ···················· | |
| D | 4960 hCoV-19/USA/CT-Yale-271/2020\|EPI_ISL_469195\|2020-0 | ···················· | ···················· | ···················· | ···················· | ···················· | ···················· | |
| D | 4961 hCoV-19/USA/CT-Yale-273/2020\|EPI_ISL_469196\|2020-0 | ···················· | ···················· | ···················· | ···················· | ···················· | ···················· | |
| D | 4962 hCoV-19/USA/CT-Yale-274/2020\|EPI_ISL_469197\|2020-0 | ···················· | ···················· | ···················· | ···················· | ···················· | ···················· | |
| D | 4963 hCoV-19/Israel/CVL-n-6105/2020\|EPI_ISL_475000\|2020 | ···················· | ···················· | ···················· | ···················· | ···················· | ···················· | |
| D | 4964 hCoV-19/Israel/CVL-n-6110/2020\|EPI_ISL_475001\|2020 | ···················· | ···················· | ···················· | ···················· | ···················· | ···················· | |
| D | 4965 hCoV-19/Israel/CVL-n-6132/2020\|EPI_ISL_475002\|2020 | ···················· | ···················· | ···················· | ···················· | ···················· | ···················· | |
| D | 4966 hCoV-19/Israel/CVL-n-6136/2020\|EPI_ISL_475003\|2020 | ···················· | ···················· | ···················· | ···················· | ···················· | ···················· | |
| D | 4967 hCoV-19/Israel/CVL-n-7201/2020\|EPI_ISL_475004\|2020 | ···················· | ···················· | ···················· | ···················· | ···················· | ···················· | |
| D | 4968 hCoV-19/Israel/CVL-n-7427/2020\|EPI_ISL_475005\|2020 | ···················· | ···················· | ···················· | ···················· | ···················· | ···················· | |
| D | 4969 hCoV-19/Israel/CVL-n-7909/2020\|EPI_ISL_475006\|2020 | ···················· | ···················· | ···················· | ···················· | ···················· | ···················· | |

| | | | | | | | | |
|---|---|---|---|---|---|---|---|---|
| D | 4970 | hCoV-19/Israel/CVL-n-7920/2020\|EPI_ISL_475007\|2020 | ···················· | ···················· | ····A··············· | ···················· | ···················· | ···················· |
| D | 4971 | hCoV-19/Israel/CVL-n-8411/2020\|EPI_ISL_475008\|2020 | ···················· | ···················· | ···················· | ···················· | ···················· | ···················· |
| D | 4972 | hCoV-19/Israel/CVL-n-12003/2020\|EPI_ISL_475009\|202 | ···················· | ···················· | ···················· | ···················· | ···················· | ···················· |
| D | 4973 | hCoV-19/Israel/CVL-n-6112/2020\|EPI_ISL_475010\|2020 | ···················· | ···················· | ···················· | ···················· | ···················· | ···················· |
| D | 4974 | hCoV-19/Israel/CVL-n-7451/2020\|EPI_ISL_475011\|2020 | ···················· | ···················· | ···················· | ···················· | ···················· | ···················· |
| D | 4975 | hCoV-19/Israel/CVL-n-7501/2020\|EPI_ISL_475012\|2020 | ···················· | ···················· | ···················· | ···················· | ···················· | ···················· |
| D | 4976 | hCoV-19/Israel/CVL-n-5908/2020\|EPI_ISL_475013\|2020 | ···················· | ···················· | ···················· | ···················· | ···················· | ···················· |
| D | 4977 | hCoV-19/Israel/CVL-n-5999/2020\|EPI_ISL_475014\|2020 | ···················· | ···················· | ···················· | ···················· | ···················· | ···················· |
| D | 4978 | hCoV-19/Israel/CVL-n-6142/2020\|EPI_ISL_475015\|2020 | ···················· | ···················· | ···················· | ···················· | ···················· | ···················· |
| D | 4979 | hCoV-19/Israel/CVL-n-10253/2020\|EPI_ISL_475016\|202 | ···················· | ···················· | ···················· | ···················· | ···················· | ···················· |
| D | 4980 | hCoV-19/Israel/CVL-n-11594/2020\|EPI_ISL_475017\|202 | ···················· | ···················· | ···················· | ···················· | ···················· | ···················· |
| D | 4981 | hCoV-19/Israel/CVL-n-5936/2020\|EPI_ISL_475018\|2020 | ···················· | ···················· | ···················· | ···················· | ···················· | ···················· |
| D | 4982 | hCoV-19/Israel/CVL-n-5991/2020\|EPI_ISL_475019\|2020 | ···················· | ···················· | ···················· | ···················· | ···················· | ···················· |
| D | 4983 | hCoV-19/Israel/CVL-n-10262/2020\|EPI_ISL_475020\|202 | ···················· | ···················· | ···················· | ···················· | ···················· | ···················· |
| D | 4984 | hCoV-19/Israel/CVL-n-6134/2020\|EPI_ISL_475021\|2020 | ···················· | ···················· | ···················· | ···················· | ···················· | ···················· |
| D | 4985 | hCoV-19/Israel/CVL-n-6157/2020\|EPI_ISL_475022\|2020 | ···················· | ···················· | ···················· | ···················· | ···················· | ···················· |
| D | 4986 | hCoV-19/Israel/CVL-n-7422/2020\|EPI_ISL_475023\|2020 | ···················· | ···················· | ···················· | ···················· | ···················· | ···················· |
| D | 4987 | hCoV-19/Israel/CVL-n-7462/2020\|EPI_ISL_475024\|2020 | ···················· | ···················· | ···················· | ···················· | ···················· | ···················· |
| D | 4988 | hCoV-19/Israel/CVL-n-7465/2020\|EPI_ISL_475025\|2020 | ···················· | ···················· | ···················· | ···················· | ···················· | ···················· |
| D | 4989 | hCoV-19/Belgium/UGent-102/2020\|EPI_ISL_475062\|2020 | ···················· | ···················· | ···················· | ···················· | ···················· | ···················· |
| D | 4994 | hCoV-19/Belgium/UGent-107/2020\|EPI_ISL_475067\|2020 | ···················· | ···················· | ·········T·········· | ···················· | ···················· | ···················· |
| D | 4996 | hCoV-19/Belgium/UGent-109/2020\|EPI_ISL_475069\|2020 | ···················· | ···················· | ···················· | ···················· | ···················· | ···················· |
| D | 4997 | hCoV-19/Brazil/SP-L2-CAMPI53/2020\|EPI_ISL_476341\|2 | ···················· | ···················· | ···················· | ···················· | ···················· | ···················· |
| D | 4998 | hCoV-19/Brazil/SP-L4-CAMPI51/2020\|EPI_ISL_476395\|2 | ···················· | ···················· | ···················· | ···················· | ···················· | ···················· |
| D | 4999 | hCoV-19/Brazil/SP-L4-CAMPI65/2020\|EPI_ISL_476398\|2 | ···················· | ···················· | ···················· | ···················· | ···················· | ···················· |
| D | 5000 | hCoV-19/Hungary/US-17911c/2020\|EPI_ISL_477615\|2020 | ···················· | ···················· | ···················· | ···················· | ···················· | ···················· |
| D | 5001 | hCoV-19/Hungary/US-18161c/2020\|EPI_ISL_477616\|2020 | ···················· | ···················· | ···················· | ···················· | ···················· | ···················· |
| D | 5002 | hCoV-19/Hungary/US-18191c/2020\|EPI_ISL_477617\|2020 | ···················· | ···················· | ···················· | ···················· | ···················· | ···················· |
| D | 5003 | hCoV-19/Hungary/US-2587w/2020\|EPI_ISL_477618\|2020- | ···················· | ···················· | ···················· | ···················· | ···················· | ···················· |
| D | 5004 | hCoV-19/Hungary/US-2823w/2020\|EPI_ISL_477619\|2020- | ···················· | ···················· | ···················· | ···················· | ···················· | ···················· |
| D | 5005 | hCoV-19/Hungary/US-2901w/2020\|EPI_ISL_477620\|2020- | ···················· | ···················· | ···················· | ···················· | ···················· | ···················· |
| D | 5006 | hCoV-19/Hungary/US-2953w/2020\|EPI_ISL_477621\|2020- | ···················· | ···················· | ···················· | ···················· | ···················· | ···················· |
| D | 5007 | hCoV-19/Hungary/US-3761w/2020\|EPI_ISL_477622\|2020- | ···················· | ···················· | ···················· | ···················· | ···················· | ···················· |
| D | 5008 | hCoV-19/Hungary/US-482w/2020\|EPI_ISL_477623\|2020-0 | ···················· | ···················· | ···················· | ···················· | ···················· | ···················· |
| D | 5009 | hCoV-19/Hungary/US-539w/2020\|EPI_ISL_477624\|2020-0 | ···················· | ···················· | ···················· | ···················· | ···················· | ···················· |
| D | 5010 | hCoV-19/Hungary/US-568w/2020\|EPI_ISL_477625\|2020-0 | ···················· | ···················· | ···················· | ···················· | ···················· | ···················· |
| D | 5011 | hCoV-19/USA/VA-DCLS-0405/2020\|EPI_ISL_477631\|2020- | ···················· | ···················· | ···················· | ···················· | ···················· | ···················· |
| D | 5012 | hCoV-19/USA/VA-DCLS-0416/2020\|EPI_ISL_477632\|2020- | ···················· | ···················· | ···················· | ···················· | ···················· | ···················· |
| D | 5013 | hCoV-19/USA/VA-DCLS-0417/2020\|EPI_ISL_477633\|2020- | ···················· | ···················· | ···················· | ···················· | ···················· | ···················· |
| D | 5014 | hCoV-19/USA/VA-DCLS-0418/2020\|EPI_ISL_477634\|2020- | ···················· | ···················· | ···················· | ···················· | ···················· | ···················· |
| D | 5015 | hCoV-19/USA/VA-DCLS-0419/2020\|EPI_ISL_477635\|2020- | ···················· | ···················· | ···················· | ···················· | ···················· | ···················· |
| D | 5016 | hCoV-19/USA/VA-DCLS-0420/2020\|EPI_ISL_477636\|2020- | ···················· | ···················· | ···················· | ···················· | ···················· | ···················· |
| D | 5017 | hCoV-19/USA/VA-DCLS-0421/2020\|EPI_ISL_477637\|2020- | ···················· | ···················· | ···················· | ···················· | ···················· | ···················· |
| D | 5018 | hCoV-19/USA/VA-DCLS-0422/2020\|EPI_ISL_477638\|2020- | ···················· | ···················· | ···················· | ···················· | ···················· | ···················· |
| D | 5019 | hCoV-19/USA/VA-DCLS-0423/2020\|EPI_ISL_477639\|2020- | ···················· | ···················· | ···················· | ···················· | ···················· | ···················· |
| D | 5020 | hCoV-19/USA/VA-DCLS-0425/2020\|EPI_ISL_477641\|2020- | ···················· | ···················· | ···················· | ···················· | ···················· | ···················· |
| D | 5021 | hCoV-19/USA/VA-DCLS-0426/2020\|EPI_ISL_477642\|2020- | ···················· | ···················· | ···················· | ···················· | ···················· | ···················· |
| D | 5022 | hCoV-19/USA/VA-DCLS-0427/2020\|EPI_ISL_477643\|2020- | ···················· | ···················· | ···················· | ···················· | ···················· | ···················· |
| D | 5023 | hCoV-19/USA/VA-DCLS-0428/2020\|EPI_ISL_477644\|2020- | ···················· | ···················· | ···················· | ···················· | ···················· | ···················· |
| D | 5024 | hCoV-19/USA/VA-DCLS-0429/2020\|EPI_ISL_477645\|2020- | ···················· | ···················· | ···················· | ···················· | ···················· | ···················· |
| D | 5025 | hCoV-19/USA/VA-DCLS-0430/2020\|EPI_ISL_477646\|2020- | ···················· | ···················· | ···················· | ···················· | ···················· | ···················· |
| D | 5026 | hCoV-19/USA/VA-DCLS-0431/2020\|EPI_ISL_477647\|2020- | ···················· | ···················· | ···················· | ···················· | ···················· | ···················· |
| D | 5027 | hCoV-19/USA/VA-DCLS-0432/2020\|EPI_ISL_477648\|2020- | ···················· | ···················· | ···················· | ···················· | ···················· | ···················· |
| D | 5028 | hCoV-19/USA/VA-DCLS-0433/2020\|EPI_ISL_477649\|2020- | ···················· | ···················· | ···················· | ···················· | ···················· | ···················· |
| D | 5029 | hCoV-19/USA/VA-DCLS-0434/2020\|EPI_ISL_477650\|2020- | ···················· | ···················· | ···················· | ···················· | ···················· | ···················· |
| D | 5030 | hCoV-19/USA/VA-DCLS-0435/2020\|EPI_ISL_477651\|2020- | ···················· | ···················· | ···················· | ···················· | ···················· | ···················· |
| D | 5031 | hCoV-19/USA/VA-DCLS-0436/2020\|EPI_ISL_477652\|2020- | ···················· | ···················· | ···················· | ···················· | ···················· | ···················· |
| D | 5032 | hCoV-19/USA/VA-DCLS-0437/2020\|EPI_ISL_477653\|2020- | ···················· | ···················· | ···················· | ···················· | ···················· | ···················· |
| D | 5033 | hCoV-19/USA/VA-DCLS-0438/2020\|EPI_ISL_477654\|2020- | ···················· | ···················· | ···················· | ···················· | ···················· | ···················· |
| D | 5034 | hCoV-19/USA/VA-DCLS-0439/2020\|EPI_ISL_477655\|2020- | ···················· | ···················· | ···················· | ···················· | ···················· | ···················· |
| D | 5035 | hCoV-19/USA/VA-DCLS-0440/2020\|EPI_ISL_477656\|2020- | ···················· | ···················· | ···················· | ···················· | ···················· | ···················· |
| D | 5036 | hCoV-19/USA/VA-DCLS-0441/2020\|EPI_ISL_477657\|2020- | ···················· | ···················· | ···················· | ···················· | ···················· | ···················· |
| D | 5037 | hCoV-19/USA/VA-DCLS-0442/2020\|EPI_ISL_477658\|2020- | ···················· | ···················· | ···················· | ···················· | ···················· | ···················· |
| D | 5038 | hCoV-19/USA/VA-DCLS-0443/2020\|EPI_ISL_477659\|2020- | ···················· | ···················· | ···················· | ···················· | ···················· | ···················· |
| D | 5039 | hCoV-19/USA/VA-DCLS-0444/2020\|EPI_ISL_477660\|2020- | ···················· | ···················· | ···················· | ···················· | ···················· | ···················· |
| D | 5040 | hCoV-19/USA/VA-DCLS-0447/2020\|EPI_ISL_477661\|2020- | ···················· | ···················· | ···················· | ···················· | ···················· | ···················· |
| D | 5041 | hCoV-19/USA/VA-DCLS-0449/2020\|EPI_ISL_477662\|2020- | ···················· | ···················· | ···················· | ···················· | ···················· | ···················· |
| D | 5042 | hCoV-19/USA/VA-DCLS-0450/2020\|EPI_ISL_477663\|2020- | ···················· | ···················· | ···················· | ···················· | ···················· | ···················· |
| D | 5043 | hCoV-19/USA/VA-DCLS-0451/2020\|EPI_ISL_477664\|2020- | ···················· | ···················· | ···················· | ···················· | ···················· | ···················· |
| D | 5044 | hCoV-19/USA/VA-DCLS-0452/2020\|EPI_ISL_477665\|2020- | ···················· | ···················· | ···················· | ···················· | ···················· | ···················· |
| D | 5045 | hCoV-19/USA/VA-DCLS-0454/2020\|EPI_ISL_477666\|2020- | ···················· | ···················· | ···················· | ···················· | ···················· | ···················· |
| D | 5046 | hCoV-19/USA/VA-DCLS-0456/2020\|EPI_ISL_477667\|2020- | ···················· | ···················· | ···················· | ···················· | ···················· | ···················· |
| D | 5047 | hCoV-19/USA/VA-DCLS-0457/2020\|EPI_ISL_477668\|2020- | ···················· | ···················· | ···················· | ···················· | ···················· | ···················· |
| D | 5048 | hCoV-19/USA/VA-DCLS-0459/2020\|EPI_ISL_477669\|2020- | ···················· | ···················· | ···················· | ···················· | ···················· | ···················· |
| D | 5049 | hCoV-19/USA/VA-DCLS-0460/2020\|EPI_ISL_477670\|2020- | ···················· | ···················· | ···················· | ···················· | ···················· | ···················· |
| D | 5050 | hCoV-19/USA/VA-DCLS-0462/2020\|EPI_ISL_477671\|2020- | ···················· | ···················· | ···················· | ···················· | ···················· | ···················· |
| D | 5051 | hCoV-19/USA/VA-DCLS-0466/2020\|EPI_ISL_477672\|2020- | ···················· | ···················· | ···················· | ···················· | ···················· | ·········A·········· |
| D | 5052 | hCoV-19/USA/CT-UW-2435/2020\|EPI_ISL_477673\|2020-03 | ···················· | ···················· | ···················· | ···················· | ···················· | ···················· |

| | | | | | | | |
|---|---|---|---|---|---|---|---|
| D | 5053 hCoV-19/USA/CT-UW-2446/2020\|EPI_ISL_477674\|2020-03 | ···················· | ···················· | ···················· | ···················· | ···················· | ···················· |
| D | 5054 hCoV-19/USA/CT-UW-2447/2020\|EPI_ISL_477675\|2020-03 | ···················· | ···················· | ···················· | ···················· | ···················· | ···················· |
| D | 5055 hCoV-19/USA/CT-UW-2466/2020\|EPI_ISL_477676\|2020-03 | ···················· | ···················· | ···················· | ···················· | ···················· | ···················· |
| D | 5056 hCoV-19/USA/CT-UW-2468/2020\|EPI_ISL_477677\|2020-03 | ···················· | ···················· | ···················· | ···················· | ···················· | ···················· |
| D | 5057 hCoV-19/USA/CT-UW-2473/2020\|EPI_ISL_477678\|2020-03 | ···················· | ···················· | ···················· | ···················· | ···················· | ···················· |
| D | 5058 hCoV-19/USA/CT-UW-2487/2020\|EPI_ISL_477679\|2020-03 | ···················· | ···················· | ···················· | ···················· | ···················· | ···················· |
| D | 5059 hCoV-19/USA/ID-UW-2047/2020\|EPI_ISL_477680\|2020-03 | ···················· | ···················· | ···················· | ···················· | ···················· | ···················· |
| D | 5060 hCoV-19/USA/ID-UW-2067/2020\|EPI_ISL_477681\|2020-03 | ···················· | ···················· | ···················· | ···················· | ···················· | ···················· |
| D | 5061 hCoV-19/USA/ID-UW-2425/2020\|EPI_ISL_477682\|2020-03 | ···················· | ···················· | ···················· | ···················· | ···················· | ···················· |
| D | 5062 hCoV-19/USA/UN-UW-1671/2020\|EPI_ISL_477683\|2020-03 | ···················· | ···················· | ···················· | ···················· | ···················· | ···················· |
| D | 5063 hCoV-19/USA/UN-UW-2476/2020\|EPI_ISL_477684\|2020-03 | ···················· | ···················· | ···················· | ···················· | ···················· | ···················· |
| D | 5064 hCoV-19/USA/WA-UW-1282/2020\|EPI_ISL_477685\|2020-03 | ···················· | ···················· | ···················· | ···················· | ···················· | ···················· |
| D | 5065 hCoV-19/USA/WA-UW-1412/2020\|EPI_ISL_477686\|2020-03 | ···················· | ···················· | ···················· | ···················· | ···················· | ···················· |
| D | 5066 hCoV-19/USA/WA-UW-1481/2020\|EPI_ISL_477687\|2020-03 | ···················· | ···················· | ···················· | ···················· | ···················· | ···················· |
| D | 5067 hCoV-19/England/OXON-B0A6B/2020\|EPI_ISL_478931\|202 | ···················· | ···················· | ···················· | ···················· | ···················· | ···················· |
| D | 5068 hCoV-19/England/OXON-B0A89/2020\|EPI_ISL_478932\|202 | ···················· | ···················· | ···················· | ···················· | ···················· | ···················· |
| D | 5069 hCoV-19/England/OXON-B0A98/2020\|EPI_ISL_478933\|202 | ···················· | ···················· | ···················· | ···················· | ···················· | ···················· |
| D | 5070 hCoV-19/England/OXON-B0AA7/2020\|EPI_ISL_478934\|202 | ···················· | ···················· | ···················· | ···················· | ···················· | ···················· |
| D | 5071 hCoV-19/England/OXON-B0AF2/2020\|EPI_ISL_478936\|202 | ···················· | ···················· | ···················· | ···················· | ···················· | ···················· |
| D | 5072 hCoV-19/England/OXON-B0B0E/2020\|EPI_ISL_478937\|202 | ···················· | ···················· | ···················· | ···················· | ···················· | ···················· |
| D | 5073 hCoV-19/England/OXON-B0B86/2020\|EPI_ISL_478939\|202 | ···················· | ···················· | ···················· | ···················· | ···················· | ···················· |
| D | 5074 hCoV-19/England/OXON-B0B95/2020\|EPI_ISL_478940\|202 | ···················· | ···················· | ···················· | ···················· | ···················· | ···················· |
| D | 5075 hCoV-19/England/OXON-B0BA4/2020\|EPI_ISL_478941\|202 | ···················· | ···················· | ···················· | ···················· | ···················· | ···················· |
| D | 5076 hCoV-19/England/OXON-B0BD1/2020\|EPI_ISL_478943\|202 | ···················· | ···················· | ···················· | ···················· | ···················· | ···················· |
| D | 5077 hCoV-19/England/OXON-B0BFF/2020\|EPI_ISL_478945\|202 | ···················· | ···················· | ···················· | ···················· | ···················· | ···················· |
| D | 5078 hCoV-19/England/OXON-B0C38/2020\|EPI_ISL_478948\|202 | ···················· | ·A·················· | ···················· | ···················· | ···················· | ···················· |
| D | 5079 hCoV-19/England/OXON-B0C56/2020\|EPI_ISL_478949\|202 | ···················· | ·A·················· | ···················· | ···················· | ···················· | ···················· |
| D | 5080 hCoV-19/England/OXON-B0C74/2020\|EPI_ISL_478950\|202 | ···················· | ···················· | ···················· | ···················· | ···················· | ···················· |
| D | 5081 hCoV-19/England/OXON-B0CB0/2020\|EPI_ISL_478952\|202 | ···················· | ···················· | ···················· | ···················· | ···················· | ···················· |
| D | 5082 hCoV-19/England/OXON-B0CDE/2020\|EPI_ISL_478954\|202 | ···················· | ···················· | ···················· | ···················· | ···················· | ···················· |
| D | 5083 hCoV-19/England/OXON-B0CED/2020\|EPI_ISL_478955\|202 | ···················· | ···················· | ···················· | ···················· | ···················· | ···················· |
| D | 5084 hCoV-19/England/OXON-B0CFC/2020\|EPI_ISL_478956\|202 | ···················· | ···················· | ···················· | ···················· | ···················· | ···················· |
| D | 5085 hCoV-19/England/OXON-B0D08/2020\|EPI_ISL_478957\|202 | ···················· | ···················· | ···················· | ···················· | ···················· | ···················· |
| D | 5086 hCoV-19/England/OXON-B0D35/2020\|EPI_ISL_478959\|202 | ···················· | ···················· | ···················· | ···················· | ···················· | ···················· |
| D | 5087 hCoV-19/England/OXON-B0D53/2020\|EPI_ISL_478960\|202 | ···················· | ···················· | ···················· | ···················· | ···················· | ···················· |
| D | 5088 hCoV-19/England/OXON-B0D62/2020\|EPI_ISL_478961\|202 | ···················· | ···················· | ···················· | ···················· | ···················· | ···················· |
| D | 5089 hCoV-19/England/OXON-B0D71/2020\|EPI_ISL_478962\|202 | ···················· | ···················· | ···················· | ···················· | ···················· | ···················· |
| D | 5090 hCoV-19/England/OXON-B0D80/2020\|EPI_ISL_478963\|202 | ···················· | ···················· | ···················· | ···················· | ···················· | ···················· |
| D | 5091 hCoV-19/England/OXON-B0DAE/2020\|EPI_ISL_478965\|202 | ···················· | ···················· | ···················· | ···················· | ···················· | ···················· |
| D | 5092 hCoV-19/England/OXON-B0E05/2020\|EPI_ISL_478967\|202 | ···················· | ···················· | ···················· | ···················· | ···················· | ···················· |
| D | 5093 hCoV-19/England/OXON-B0E14/2020\|EPI_ISL_478968\|202 | ···················· | ···················· | ···················· | ···················· | ···················· | ···················· |
| D | 5094 hCoV-19/England/OXON-B0E50/2020\|EPI_ISL_478969\|202 | ···················· | ···················· | ···················· | ···················· | ···················· | ···················· |
| D | 5095 hCoV-19/England/OXON-B0EAB/2020\|EPI_ISL_478973\|202 | ···················· | ···················· | ···················· | ···················· | ···················· | ···················· |
| D | 5096 hCoV-19/England/OXON-B0EBA/2020\|EPI_ISL_478974\|202 | ···················· | ···················· | ···················· | ···················· | ···················· | ···················· |
| D | 5097 hCoV-19/England/OXON-B0ED8/2020\|EPI_ISL_478975\|202 | ···················· | ···················· | ···················· | ···················· | ···················· | ···················· |
| D | 5098 hCoV-19/England/OXON-B0EF6/2020\|EPI_ISL_478976\|202 | ···················· | ···················· | ···················· | ···················· | ···················· | ···················· |
| D | 5099 hCoV-19/England/OXON-B0F11/2020\|EPI_ISL_478977\|202 | ···················· | ···················· | ···················· | ···················· | ···················· | ···················· |
| D | 5100 hCoV-19/England/OXON-B0F3F/2020\|EPI_ISL_478979\|202 | ···················· | ···················· | ···················· | ···················· | ···················· | ···················· |
| D | 5101 hCoV-19/England/OXON-B0F5D/2020\|EPI_ISL_478980\|202 | ···················· | ···················· | ···················· | ···················· | ···················· | ···················· |
| D | 5102 hCoV-19/England/OXON-B0F6C/2020\|EPI_ISL_478981\|202 | ···················· | ···················· | ···················· | ···················· | ···················· | ···················· |
| D | 5103 hCoV-19/England/OXON-B0F7B/2020\|EPI_ISL_478982\|202 | ···················· | ···················· | ···················· | ···················· | ···················· | ···················· |
| D | 5104 hCoV-19/England/OXON-B0F8A/2020\|EPI_ISL_478983\|202 | ···················· | ···················· | ···················· | ···················· | ···················· | ···················· |
| D | 5105 hCoV-19/England/OXON-B0FB7/2020\|EPI_ISL_478984\|202 | ···················· | ···················· | ···················· | ···················· | ···················· | ···················· |
| D | 5106 hCoV-19/England/OXON-B0FE4/2020\|EPI_ISL_478985\|202 | ···················· | ···················· | ···················· | ···················· | ···················· | ···················· |
| D | 5107 hCoV-19/England/OXON-B0FF3/2020\|EPI_ISL_478986\|202 | ···················· | ···················· | ···················· | ···················· | ···················· | ···················· |
| D | 5108 hCoV-19/England/OXON-B101D/2020\|EPI_ISL_478987\|202 | ···················· | ···················· | ···················· | ···················· | ···················· | ···················· |
| D | 5109 hCoV-19/England/OXON-B102C/2020\|EPI_ISL_478988\|202 | ···················· | ···················· | ···················· | ···················· | ···················· | ···················· |
| D | 5110 hCoV-19/England/OXON-B103B/2020\|EPI_ISL_478989\|202 | ···················· | ···················· | ···················· | ···················· | ···················· | ···················· |
| D | 5111 hCoV-19/England/OXON-B1068/2020\|EPI_ISL_478990\|202 | ···················· | ···················· | ···················· | ···················· | ···················· | ···················· |
| D | 5112 hCoV-19/England/OXON-B1086/2020\|EPI_ISL_478991\|202 | ···················· | ···················· | ···················· | ···················· | ···················· | ···················· |
| D | 5113 hCoV-19/England/OXON-B1095/2020\|EPI_ISL_478992\|202 | ···················· | ···················· | ···················· | ···················· | ···················· | ···················· |
| D | 5114 hCoV-19/England/OXON-B10A4/2020\|EPI_ISL_478993\|202 | ···················· | ···················· | ···················· | ···················· | ···················· | ···················· |
| D | 5115 hCoV-19/England/OXON-B10B3/2020\|EPI_ISL_478994\|202 | ···················· | ···················· | ···················· | ···················· | ···················· | ···················· |
| D | 5116 hCoV-19/England/OXON-B10C2/2020\|EPI_ISL_478995\|202 | ···················· | ···················· | ···················· | ···················· | ···················· | ···················· |
| D | 5117 hCoV-19/England/OXON-B10E0/2020\|EPI_ISL_478996\|202 | ···················· | ···················· | ···················· | ···················· | ···················· | ···················· |
| D | 5118 hCoV-19/England/OXON-B110B/2020\|EPI_ISL_478997\|202 | ···················· | ···················· | ···················· | ···················· | ···················· | ···················· |
| D | 5119 hCoV-19/England/OXON-B1147/2020\|EPI_ISL_478998\|202 | ···················· | ···········A········ | ···················· | ···················· | ···················· | ···················· |
| D | 5120 hCoV-19/England/OXON-B1165/2020\|EPI_ISL_478999\|202 | ···················· | ···················· | ···················· | ···················· | ···················· | ···················· |
| D | 5121 hCoV-19/England/201021655/2020\|EPI_ISL_464300\|2020 | ···················· | ···················· | ···················· | ···················· | ···················· | ···················· |
| D | 5124 hCoV-19/England/20104003304/2020\|EPI_ISL_464307\|20 | ···················· | ···················· | ···················· | ···················· | ···················· | ···················· |
| D | 5125 hCoV-19/England/201040036/2020\|EPI_ISL_464310\|2020 | ···················· | ···················· | ···················· | ···················· | ···················· | ···················· |
| D | 5126 hCoV-19/England/201040041/2020\|EPI_ISL_464311\|2020 | ···················· | ···················· | ···················· | ···················· | ···················· | ···················· |
| D | 5128 hCoV-19/England/20104004304/2020\|EPI_ISL_464314\|20 | ···················· | ···················· | ···················· | ···················· | ···················· | ···················· |
| D | 5130 hCoV-19/England/201040073/2020\|EPI_ISL_464318\|2020 | ···················· | ···················· | ···················· | ···················· | ···················· | ···················· |
| D | 5132 hCoV-19/England/201040079/2020\|EPI_ISL_464323\|2020 | ···················· | ···················· | ···················· | ···················· | ···················· | ···················· |
| D | 5133 hCoV-19/England/20194157001/2020\|EPI_ISL_465621\|20 | ···················· | ···················· | ···················· | ···················· | ···················· | ···················· |
| D | 5134 hCoV-19/England/20196026804/2020\|EPI_ISL_465623\|20 | ···················· | ···················· | ···················· | ···················· | ···················· | ···················· |
| D | 5135 hCoV-19/England/20196064904/2020\|EPI_ISL_465626\|20 | ···················· | ···················· | ···················· | ···················· | ···················· | ···················· |

| | | | | | | | | |
|---|---|---|---|---|---|---|---|---|
| D | 5136 | hCoV-19/England/20196112204/2020\|EPI_ISL_465628\|20 | ···················· | ···················· | ···················· | ···················· | ···················· | ···················· |
| D | 5137 | hCoV-19/England/20198006204/2020\|EPI_ISL_465633\|20 | ···················· | ···················· | ···················· | ···················· | ···················· | ···················· |
| D | 5138 | hCoV-19/England/20198121604/2020\|EPI_ISL_465640\|20 | ···················· | ···················· | ···················· | ···················· | ···················· | ···················· |
| D | 5140 | hCoV-19/England/20204113704/2020\|EPI_ISL_465652\|20 | ···················· | ···················· | ···················· | ···················· | ···················· | ···················· |
| D | 5141 | hCoV-19/England/20206069204/2020\|EPI_ISL_465661\|20 | ···················· | ···················· | ···················· | ···················· | ···················· | ···················· |
| D | 5142 | hCoV-19/England/20206138304/2020\|EPI_ISL_465662\|20 | ···················· | ···················· | ···················· | ···················· | ···················· | ···················· |
| D | 5143 | hCoV-19/England/20206138804/2020\|EPI_ISL_465663\|20 | ···················· | ···················· | ···················· | ···················· | ···················· | ···················· |
| D | 5144 | hCoV-19/England/20206139404/2020\|EPI_ISL_465664\|20 | ···················· | ···················· | ···················· | ···················· | ···················· | ···················· |
| D | 5145 | hCoV-19/Germany/BY-MVP-0244/2020\|EPI_ISL_466900\|20 | ···················· | ···················· | ···················· | ···················· | ···················· | ···················· |
| D | 5146 | hCoV-19/Germany/BY-MVP-0246/2020\|EPI_ISL_466901\|20 | ···················· | ···················· | ···················· | ···················· | ···················· | ···················· |
| D | 5147 | hCoV-19/Germany/BY-MVP-0247/2020\|EPI_ISL_466902\|20 | ···················· | ···················· | ···················· | ···················· | ···················· | ···················· |
| D | 5148 | hCoV-19/Germany/BY-MVP-0249/2020\|EPI_ISL_466903\|20 | ···················· | ···················· | ···················· | ···················· | ···················· | ···················· |
| D | 5149 | hCoV-19/Germany/BY-MVP-0250/2020\|EPI_ISL_466904\|20 | ···················· | ···················· | ···················· | ···················· | ···················· | ···················· |
| D | 5150 | hCoV-19/Germany/BY-MVP-0251/2020\|EPI_ISL_466905\|20 | ···················· | ···················· | ···················· | ···················· | ···················· | ···················· |
| D | 5151 | hCoV-19/Germany/BY-MVP-0253/2020\|EPI_ISL_466906\|20 | ···················· | ···················· | ···················· | ···················· | ···················· | ···················· |
| D | 5152 | hCoV-19/Germany/BY-MVP-0254/2020\|EPI_ISL_466907\|20 | ···················· | ···················· | ···················· | ···················· | ···················· | ···················· |
| D | 5153 | hCoV-19/Germany/BY-MVP-0283/2020\|EPI_ISL_466909\|20 | ···················· | ···················· | ···················· | ···················· | ···················· | ···················· |
| D | 5154 | hCoV-19/Germany/BY-MVP-0268/2020\|EPI_ISL_466910\|20 | ···················· | ···················· | ···················· | ···················· | ···················· | ···················· |
| D | 5158 | hCoV-19/Switzerland/BS-ETHZ-100029/2020\|EPI_ISL_46 | ···················· | ···················· | ···················· | ···················· | ···················· | ···················· |
| D | 5160 | hCoV-19/Switzerland/BE-ETHZ-100031/2020\|EPI_ISL_46 | ···················· | ···················· | ···················· | ···················· | ···················· | ···················· |
| D | 5163 | hCoV-19/Switzerland/BE-ETHZ-100035/2020\|EPI_ISL_46 | ···················· | ···················· | ···················· | ···················· | ···················· | ···················· |
| D | 5164 | hCoV-19/Switzerland/BS-ETHZ-100036/2020\|EPI_ISL_46 | ···················· | ···················· | ···················· | ···················· | ···················· | ···················· |
| D | 5165 | hCoV-19/Switzerland/VD-ETHZ-100037/2020\|EPI_ISL_46 | ···················· | ···················· | ···················· | ···················· | ······S············· | ···················· |
| D | 5173 | hCoV-19/Switzerland/TI-ETHZ-100048/2020\|EPI_ISL_46 | ···················· | ···················· | ···················· | ···················· | ···················· | ···················· |
| D | 5181 | hCoV-19/Switzerland/ZH-ETHZ-100057/2020\|EPI_ISL_46 | ···················· | ·A·················· | ···················· | ···················· | ···················· | ···················· |
| D | 5182 | hCoV-19/Switzerland/VD-ETHZ-100058/2020\|EPI_ISL_46 | ···················· | ·A·················· | ···················· | ···················· | ······S············· | ···················· |
| D | 5193 | hCoV-19/Switzerland/TI-ETHZ-100069/2020\|EPI_ISL_46 | ···················· | ···················· | ···················· | ···················· | ···················· | ···················· |
| D | 5195 | hCoV-19/Switzerland/TI-ETHZ-100073/2020\|EPI_ISL_46 | ···················· | ···················· | ···················· | ···················· | ···················· | ···················· |
| D | 5196 | hCoV-19/Singapore/353/2020\|EPI_ISL_469108\|2020-05- | ···················· | ···················· | ···················· | ···················· | ···················· | ···················· |
| D | 5197 | hCoV-19/Singapore/354/2020\|EPI_ISL_469109\|2020-04- | ···················· | ···················· | ···················· | ···················· | ···················· | ···················· |
| D | 5198 | hCoV-19/Singapore/355/2020\|EPI_ISL_469110\|2020-04- | ···················· | ···················· | ···················· | ···················· | ···················· | ···················· |
| D | 5199 | hCoV-19/Singapore/356/2020\|EPI_ISL_469111\|2020-04- | ···················· | ···················· | ···················· | ···················· | ···················· | ···················· |
| D | 5200 | hCoV-19/Singapore/357/2020\|EPI_ISL_469112\|2020-04- | ···················· | ···················· | ···················· | ···················· | ···················· | ···················· |
| D | 5201 | hCoV-19/Singapore/358/2020\|EPI_ISL_469113\|2020-04- | ···················· | ···················· | ···················· | ···················· | ···················· | ··············R····· |
| D | 5202 | hCoV-19/Singapore/359/2020\|EPI_ISL_469114\|2020-04- | ···················· | ···················· | ···················· | ···················· | ···················· | ···················· |
| D | 5203 | hCoV-19/Singapore/360/2020\|EPI_ISL_469115\|2020-05- | ···················· | ···················· | ···················· | ···················· | ···················· | ···················· |
| D | 5204 | hCoV-19/Singapore/361/2020\|EPI_ISL_469116\|2020 | ···················· | ···················· | ···················· | ···················· | ···················· | ···················· |
| D | 5205 | hCoV-19/Singapore/362/2020\|EPI_ISL_469117\|2020-04- | ···················· | ···················· | ···················· | ···················· | ···················· | ···················· |
| D | 5206 | hCoV-19/Singapore/363/2020\|EPI_ISL_469118\|2020-04- | ···················· | ···················· | ···················· | ···················· | ···················· | ···················· |
| D | 5207 | hCoV-19/Singapore/364/2020\|EPI_ISL_469119\|2020-04- | ···················· | ···················· | ···················· | ···················· | ···················· | ···················· |
| D | 5208 | hCoV-19/Singapore/365/2020\|EPI_ISL_469120\|2020-04- | ···················· | ···················· | ···················· | ···················· | ···················· | ···················· |
| D | 5209 | hCoV-19/Singapore/366/2020\|EPI_ISL_469121\|2020-03- | ···················· | ···················· | ···················· | ···················· | ···················· | ···················· |
| D | 5210 | hCoV-19/Singapore/375/2020\|EPI_ISL_469124\|2020-03- | ···················· | ···················· | ···················· | ···················· | ···················· | ···················· |
| D | 5212 | hCoV-19/Singapore/377/2020\|EPI_ISL_469126\|2020-04- | ···················· | ···················· | ···················· | ···················· | ···················· | ···················· |
| D | 5213 | hCoV-19/Singapore/378/2020\|EPI_ISL_469127\|2020-03- | ···················· | ···················· | ···················· | ···················· | ···················· | ···················· |
| D | 5215 | hCoV-19/Singapore/380/2020\|EPI_ISL_469129\|2020-04- | ···················· | ···················· | ···················· | ···················· | ···················· | ···················· |
| D | 5216 | hCoV-19/Singapore/381/2020\|EPI_ISL_469130\|2020-04- | ···················· | ···················· | ···················· | ···················· | ···················· | ···················· |
| D | 5217 | hCoV-19/Singapore/382/2020\|EPI_ISL_469131\|2020-04- | ···················· | ···················· | ···················· | ···················· | ···················· | ···················· |
| D | 5218 | hCoV-19/Singapore/383/2020\|EPI_ISL_469132\|2020-02- | ···················· | ···················· | ···················· | ···················· | ···················· | ···················· |
| D | 5219 | hCoV-19/Singapore/384/2020\|EPI_ISL_469133\|2020-04- | ···················· | ···················· | ···················· | ···················· | ···················· | ···················· |
| D | 5220 | hCoV-19/Singapore/385/2020\|EPI_ISL_469134\|2020-04- | ···················· | ···················· | ···················· | ···················· | ···················· | ···················· |
| D | 5221 | hCoV-19/Singapore/386/2020\|EPI_ISL_469135\|2020-04- | ···················· | ···················· | ···················· | ···················· | ···················· | ···················· |
| D | 5222 | hCoV-19/Singapore/387/2020\|EPI_ISL_469136\|2020-04- | ···················· | ···················· | ···················· | ···················· | ···················· | ···················· |
| D | 5223 | hCoV-19/Singapore/388/2020\|EPI_ISL_469137\|2020-04- | ···················· | ···················· | ···················· | ···················· | ···················· | ···················· |
| D | 5224 | hCoV-19/Singapore/389/2020\|EPI_ISL_469138\|2020-04- | ···················· | ···················· | ···················· | ···················· | ···················· | ···················· |
| D | 5225 | hCoV-19/Singapore/390/2020\|EPI_ISL_469139\|2020-06- | ···················· | ···················· | ···················· | ···················· | ···················· | ···················· |
| D | 5226 | hCoV-19/Singapore/391/2020\|EPI_ISL_469140\|2020-04- | ···················· | ···················· | ···················· | ···················· | ···················· | ···················· |
| D | 5227 | hCoV-19/Singapore/392/2020\|EPI_ISL_469141\|2020-04- | ···················· | ···················· | ···················· | ···················· | ···················· | ···················· |
| D | 5228 | hCoV-19/Singapore/393/2020\|EPI_ISL_469142\|2020-04- | ···················· | ···················· | ···················· | ···················· | ···················· | ···················· |
| D | 5229 | hCoV-19/Singapore/394/2020\|EPI_ISL_469143\|2020-03- | ···················· | ···················· | ···················· | ···················· | ···················· | ···················· |
| D | 5230 | hCoV-19/Singapore/395/2020\|EPI_ISL_469144\|2020-04- | ···················· | ···················· | ···················· | ···················· | ···················· | ···················· |
| D | 5231 | hCoV-19/Singapore/396/2020\|EPI_ISL_469145\|2020-04- | ···················· | ···················· | ···················· | ···················· | ···················· | ···················· |
| D | 5232 | hCoV-19/Singapore/397/2020\|EPI_ISL_469146\|2020-06- | ···················· | ···················· | ···················· | ···················· | ···················· | ···················· |
| D | 5233 | hCoV-19/Singapore/398/2020\|EPI_ISL_469147\|2020-05- | ···················· | ···················· | ···················· | ···················· | ···················· | ···················· |
| D | 5234 | hCoV-19/Singapore/399/2020\|EPI_ISL_469148\|2020-04- | ···················· | ···················· | ···················· | ···················· | ···················· | ···················· |
| D | 5235 | hCoV-19/Singapore/400/2020\|EPI_ISL_469149\|2020-04- | ···················· | ···················· | ···················· | ···················· | ···················· | ···················· |
| D | 5236 | hCoV-19/USA/CT-Yale-245/2020\|EPI_ISL_469173\|2020-0 | ···················· | ···················· | ···················· | ···················· | ···················· | ···················· |
| D | 5237 | hCoV-19/USA/CT-Yale-247/2020\|EPI_ISL_469175\|2020-0 | ···················· | ···················· | ···················· | ···················· | ···················· | ···················· |
| D | 5238 | hCoV-19/USA/CT-Yale-250/2020\|EPI_ISL_469177\|2020-0 | ···················· | ···················· | ···················· | ···················· | ···················· | ···················· |
| D | 5239 | hCoV-19/USA/CT-Yale-253/2020\|EPI_ISL_469179\|2020-0 | ···················· | ···················· | ···················· | ···················· | ···················· | ···················· |
| D | 5240 | hCoV-19/USA/CT-Yale-255/2020\|EPI_ISL_469181\|2020-0 | ···················· | ···················· | ···················· | ···················· | ···················· | ···················· |
| D | 5241 | hCoV-19/USA/CT-Yale-256/2020\|EPI_ISL_469182\|2020-0 | ···················· | ···················· | ···················· | ···················· | ···················· | ···················· |
| D | 5242 | hCoV-19/USA/CT-Yale-257/2020\|EPI_ISL_469183\|2020-0 | ···················· | ···················· | ···················· | ···················· | ···················· | ···················· |
| D | 5243 | hCoV-19/USA/CT-Yale-258/2020\|EPI_ISL_469184\|2020-0 | ···················· | ···················· | ···················· | ···················· | ···················· | ···················· |
| D | 5244 | hCoV-19/USA/CT-Yale-260/2020\|EPI_ISL_469186\|2020-0 | ···················· | ···················· | ···················· | ···················· | ···················· | ···················· |
| D | 5245 | hCoV-19/USA/CT-Yale-261/2020\|EPI_ISL_469187\|2020-0 | ···················· | ···················· | ···················· | ···················· | ···················· | ···················· |
| D | 5246 | hCoV-19/USA/CT-Yale-263/2020\|EPI_ISL_469189\|2020-0 | ···················· | ···················· | ···················· | ···················· | ···················· | ···················· |
| D | 5247 | hCoV-19/England/CAMB-1AAD00/2020\|EPI_ISL_470101\|20 | ···················· | ·············A······ | ···················· | ···················· | ···················· | ···················· |

| | | | | | | | |
|---|---|---|---|---|---|---|---|
| D | 5248 hCoV-19/England/CAMB-1AA8B4/2020\|EPI_ISL_470102\|20 | ···················· | ···················· | ···················· | ···················· | ···················· | ···················· |
| D | 5249 hCoV-19/England/CAMB-1AA8F0/2020\|EPI_ISL_470103\|20 | ···················· | ···················· | ···················· | ···················· | ···················· | ···················· |
| D | 5250 hCoV-19/England/CAMB-1AABE8/2020\|EPI_ISL_470104\|20 | ···················· | ···················· | ···················· | ···················· | ···················· | ···················· |
| D | 5251 hCoV-19/England/CAMB-1AAC12/2020\|EPI_ISL_470105\|20 | ···················· | ···················· | ···················· | ···················· | ···················· | ···················· |
| D | 5252 hCoV-19/England/CAMB-1AB75C/2020\|EPI_ISL_470107\|20 | ···················· | ···················· | ···················· | ···················· | ···················· | ···················· |
| D | 5253 hCoV-19/England/CAMB-1AA9C0/2020\|EPI_ISL_470108\|20 | ···················· | ···················· | ···················· | ···················· | ···················· | ···················· |
| D | 5254 hCoV-19/England/CAMB-1AACD6/2020\|EPI_ISL_470109\|20 | ···················· | ···················· | ···················· | ···················· | ···················· | ···················· |
| D | 5255 hCoV-19/England/CAMB-1AB83B/2020\|EPI_ISL_470110\|20 | ···················· | ···················· | ···················· | ···················· | ···················· | ···················· |
| D | 5256 hCoV-19/England/CAMB-1AA6AB/2020\|EPI_ISL_470114\|20 | ···················· | ···················· | ···················· | ···················· | ···················· | ···················· |
| D | 5257 hCoV-19/England/CAMB-1AAC6D/2020\|EPI_ISL_470115\|20 | ···················· | ···················· | ···················· | ···················· | ···················· | ···················· |
| D | 5258 hCoV-19/England/CAMB-1AAC4F/2020\|EPI_ISL_470116\|20 | ···················· | ···················· | ···················· | ···················· | ···················· | ···················· |
| D | 5259 hCoV-19/England/CAMB-1ABA08/2020\|EPI_ISL_470131\|20 | ···················· | ···················· | ···················· | ···················· | ···················· | ···················· |
| D | 5260 hCoV-19/England/CAMB-1AB6F5/2020\|EPI_ISL_470132\|20 | ···················· | ···················· | ···················· | ···················· | ···················· | ···················· |
| D | 5261 hCoV-19/England/CAMB-1AA702/2020\|EPI_ISL_470133\|20 | ···················· | ···················· | ···················· | ···················· | ···················· | ···················· |
| D | 5262 hCoV-19/England/CAMB-1ABC11/2020\|EPI_ISL_470136\|20 | ···················· | ···················· | ···················· | ···················· | ···················· | ···················· |
| D | 5263 hCoV-19/England/CAMB-1ABC02/2020\|EPI_ISL_470137\|20 | ···················· | ···················· | ···················· | ···················· | ···················· | ···················· |
| D | 5264 hCoV-19/England/CAMB-1AB9CF/2020\|EPI_ISL_470140\|20 | ···················· | ···················· | ···················· | ···················· | ···················· | ···················· |
| D | 5265 hCoV-19/England/CAMB-1AAC9A/2020\|EPI_ISL_470141\|20 | ···················· | ···················· | ···················· | ···················· | ···················· | ···················· |
| D | 5266 hCoV-19/England/CAMB-1ABAAE/2020\|EPI_ISL_470142\|20 | ···················· | ···················· | ···················· | ···················· | ···················· | ···················· |
| D | 5267 hCoV-19/England/CAMB-1ABD4B/2020\|EPI_ISL_470143\|20 | ···················· | ···················· | ···················· | ···················· | ···················· | ···················· |
| D | 5268 hCoV-19/England/CAMB-1AAB24/2020\|EPI_ISL_470144\|20 | ···················· | ···················· | ···················· | ···················· | ···················· | ···················· |
| D | 5269 hCoV-19/England/CAMB-1AB9ED/2020\|EPI_ISL_470146\|20 | ···················· | ···················· | ···················· | ···················· | ···················· | ···················· |
| D | 5270 hCoV-19/England/CAMB-1AA878/2020\|EPI_ISL_470147\|20 | ···················· | ···················· | ···················· | ···················· | ···················· | ···················· |
| D | 5271 hCoV-19/England/CAMB-1ABB9C/2020\|EPI_ISL_470148\|20 | ···················· | ···················· | ···················· | ···················· | ···················· | ···················· |
| D | 5272 hCoV-19/England/CAMB-1AB68C/2020\|EPI_ISL_470150\|20 | ···················· | ···················· | ···················· | ···················· | ···················· | ···················· |
| D | 5273 hCoV-19/England/CAMB-1ABA62/2020\|EPI_ISL_470152\|20 | ···················· | ···················· | ···················· | ···················· | ···················· | ···················· |
| D | 5274 hCoV-19/England/CAMB-1AA83C/2020\|EPI_ISL_470153\|20 | ···················· | ···················· | ···················· | ···················· | ···················· | ···················· |
| D | 5275 hCoV-19/England/CAMB-1AB8A4/2020\|EPI_ISL_470154\|20 | ···················· | ···················· | ···················· | ···················· | ···················· | ···················· |
| D | 5276 hCoV-19/England/CAMB-1AABAC/2020\|EPI_ISL_470155\|20 | ···················· | ···················· | ···················· | ···················· | ···················· | ···················· |
| D | 5277 hCoV-19/England/CAMB-1AB32B/2020\|EPI_ISL_470156\|20 | ···················· | ···················· | ···················· | ···················· | ······T············· | ···················· |
| D | 5278 hCoV-19/England/CAMB-1AA5AE/2020\|EPI_ISL_470158\|20 | ···················· | ···················· | ···················· | ···················· | ···················· | ···················· |
| D | 5279 hCoV-19/England/CAMB-1AB349/2020\|EPI_ISL_470159\|20 | ···················· | ···················· | ···················· | ···················· | ···················· | ···················· |
| D | 5280 hCoV-19/England/CAMB-1AB938/2020\|EPI_ISL_470160\|20 | ···················· | ···················· | ···················· | ···················· | ···················· | ···················· |
| D | 5281 hCoV-19/England/CAMB-1AB376/2020\|EPI_ISL_470161\|20 | ···················· | ···················· | ···················· | ···················· | ···················· | ···················· |
| D | 5282 hCoV-19/England/CAMB-1ABBE7/2020\|EPI_ISL_470163\|20 | ···················· | ···················· | ···················· | ···················· | ···················· | ···················· |
| D | 5283 hCoV-19/England/CAMB-1AA80F/2020\|EPI_ISL_470166\|20 | ···················· | ···················· | ···················· | ···················· | ···················· | ···················· |
| D | 5284 hCoV-19/England/CAMB-1AA605/2020\|EPI_ISL_470167\|20 | ···················· | ···················· | ···················· | ···················· | ···················· | ···················· |
| D | 5285 hCoV-19/England/CAMB-1AB631/2020\|EPI_ISL_470170\|20 | ···················· | ···················· | ···················· | ···················· | ···················· | ···················· |
| D | 5286 hCoV-19/England/CAMB-1AA975/2020\|EPI_ISL_470171\|20 | ···················· | ···················· | ···················· | ···················· | ···················· | ···················· |
| D | 5287 hCoV-19/England/CAMB-1ABD3C/2020\|EPI_ISL_470172\|20 | ···················· | ···················· | ···················· | ···················· | ···················· | ···················· |
| D | 5288 hCoV-19/England/CAMB-1AB6C8/2020\|EPI_ISL_470173\|20 | ···················· | ···················· | ···················· | ···················· | ···················· | ···················· |
| D | 5289 hCoV-19/England/CAMB-1AB90B/2020\|EPI_ISL_470174\|20 | ···················· | ···················· | ···················· | ···················· | ···················· | ···················· |
| D | 5290 hCoV-19/England/CAMB-1ABB7E/2020\|EPI_ISL_470181\|20 | ···················· | ···················· | ···················· | ···················· | ···················· | ···················· |
| D | 5291 hCoV-19/England/CAMB-1AA76C/2020\|EPI_ISL_470182\|20 | ···················· | ···················· | ···················· | ···················· | ···················· | ···················· |
| D | 5292 hCoV-19/England/CAMB-1AB534/2020\|EPI_ISL_470183\|20 | ···················· | ···················· | ···················· | ···················· | ···················· | ···················· |
| D | 5293 hCoV-19/England/CAMB-1AB73E/2020\|EPI_ISL_470184\|20 | ···················· | ···················· | ···················· | ···················· | ···················· | ···················· |
| D | 5294 hCoV-19/England/CAMB-1AABBB/2020\|EPI_ISL_470185\|20 | ···················· | ···················· | ···················· | ···················· | ···················· | ···················· |
| D | 5295 hCoV-19/England/CAMB-1ABBF6/2020\|EPI_ISL_470186\|20 | ···················· | ···················· | ···················· | ···················· | ···················· | ···················· |
| D | 5296 hCoV-19/England/CAMB-1AAC5E/2020\|EPI_ISL_470187\|20 | ···················· | ···················· | ···················· | ···················· | ···················· | ···················· |
| D | 5297 hCoV-19/England/CAMB-1AA7B7/2020\|EPI_ISL_470188\|20 | ···················· | ···················· | ···················· | ···················· | ···················· | ···················· |
| D | 5298 hCoV-19/England/CAMB-1AABD9/2020\|EPI_ISL_470190\|20 | ···················· | ···················· | ···················· | ···················· | ···················· | ···················· |
| D | 5299 hCoV-19/England/CAMB-1ABD1E/2020\|EPI_ISL_470192\|20 | ···················· | ···················· | ···················· | ···················· | ···················· | ···················· |
| D | 5300 hCoV-19/England/CAMB-1AB992/2020\|EPI_ISL_470194\|20 | ···················· | ···················· | ···················· | ···················· | ···················· | ···················· |
| D | 5301 hCoV-19/England/CAMB-1AB8E0/2020\|EPI_ISL_470197\|20 | ···················· | ···················· | ···················· | ···················· | ···················· | ···················· |
| D | 5302 hCoV-19/England/CAMB-1AB9B0/2020\|EPI_ISL_470198\|20 | ···················· | ···················· | ···················· | ···················· | ···················· | ···················· |
| D | 5303 hCoV-19/Romania/Bucuresti-293034/2020\|EPI_ISL_4714 | ···················· | ···················· | ···················· | ···················· | ···················· | ···················· |
| D | 5304 hCoV-19/Romania/Bucuresti-294285/2020\|EPI_ISL_4714 | ···················· | ·A·················· | ···················· | ···················· | ···················· | ···················· |
| D | 5305 hCoV-19/Romania/Buzau-292638/2020\|EPI_ISL_471423\|2 | ···················· | ·A·················· | ···················· | ···················· | ···················· | ···················· |
| D | 5306 hCoV-19/Romania/Valcea/2020\|EPI_ISL_471424\|2020-06 | ···················· | ···················· | ···················· | ···················· | ···················· | ···················· |
| D | 5307 hCoV-19/South Korea/KCDC2020/2020\|EPI_ISL_471425\|2 | ···················· | ···················· | ···················· | ···················· | ···················· | ···················· |
| D | 5308 hCoV-19/South Korea/KCDC2021/2020\|EPI_ISL_471426\|2 | ···················· | ···················· | ···················· | ···················· | ···················· | ···················· |
| D | 5309 hCoV-19/Belgium/ULG-10192/2020\|EPI_ISL_471427\|2020 | ···················· | ···················· | ···················· | ···················· | ···················· | ···················· |
| D | 5310 hCoV-19/Belgium/ULG-10194/2020\|EPI_ISL_471429\|2020 | ···················· | ···················· | ···················· | ···················· | ·T·················· | ···················· |
| D | 5311 hCoV-19/Belgium/ULG-10195/2020\|EPI_ISL_471430\|2020 | ···················· | ···················· | ···················· | ···················· | ···················· | ···················· |
| D | 5312 hCoV-19/Belgium/ULG-10196/2020\|EPI_ISL_471431\|2020 | ···················· | ···················· | ···················· | ···················· | ···················· | ···················· |
| D | 5313 hCoV-19/Belgium/ULG-10200/2020\|EPI_ISL_471435\|2020 | ···················· | ···················· | ···················· | ···················· | ···················· | ···················· |
| D | 5314 hCoV-19/Belgium/ULG-10202/2020\|EPI_ISL_471436\|2020 | ···················· | ···················· | ···················· | ···················· | ···················· | ···················· |
| D | 5315 hCoV-19/South Korea/KCDC2022/2020\|EPI_ISL_471438\|2 | ···················· | ···················· | ···················· | ···················· | ···················· | ···················· |
| D | 5316 hCoV-19/South Korea/KCDC2023/2020\|EPI_ISL_471439\|2 | ···················· | ···················· | ···················· | ···················· | ···················· | ···················· |
| D | 5317 hCoV-19/South Korea/KCDC2024/2020\|EPI_ISL_471440\|2 | ···················· | ···················· | ···················· | ···················· | ···················· | ···················· |
| D | 5318 hCoV-19/South Korea/KCDC2025/2020\|EPI_ISL_471441\|2 | ···················· | ···················· | ···················· | ···················· | ···················· | ···················· |
| D | 5319 hCoV-19/South Korea/KCDC2026/2020\|EPI_ISL_471442\|2 | ···················· | ···················· | ···················· | ···················· | ···················· | ···················· |
| D | 5320 hCoV-19/South Korea/KCDC2028/2020\|EPI_ISL_471444\|2 | ···················· | ···················· | ···················· | ···················· | ···················· | ···················· |
| D | 5321 hCoV-19/South Korea/KCDC2029/2020\|EPI_ISL_471445\|2 | ···················· | ···················· | ···················· | ···················· | ···················· | ···················· |
| D | 5322 hCoV-19/South Korea/KCDC2040/2020\|EPI_ISL_471446\|2 | ···················· | ···················· | ···················· | ···················· | ···················· | ···················· |
| D | 5323 hCoV-19/South Korea/KCDC2041/2020\|EPI_ISL_471447\|2 | ···················· | ···················· | ···················· | ···················· | ···················· | ···················· |
| D | 5324 hCoV-19/Spain/CT-IrsiCaixa-JP/2020\|EPI_ISL_471472\| | ···················· | ···················· | ···················· | ···················· | ···················· | ···················· |
| D | 5325 hCoV-19/Wales/PHWC-15D8FC/2020\|EPI_ISL_472750\|2020 | ···················· | ···················· | ···················· | ···················· | ···················· | ···················· |

| | | | | | | | | | |
|---|---|---|---|---|---|---|---|---|---|
| D | 5326 | hCoV-19/Wales/PHWC-15D917/2020\|EPI_ISL_472751\|2020 | .................... | .................... | .................... | .................... | .................... | .................... | |
| D | 5327 | hCoV-19/Wales/PHWC-15D9CC/2020\|EPI_ISL_472753\|2020 | .................... | .................... | .................... | .................... | .................... | .................... | |
| D | 5328 | hCoV-19/Wales/PHWC-15DA14/2020\|EPI_ISL_472754\|2020 | .................... | .................... | .................... | .................... | .................... | .................... | |
| D | 5329 | hCoV-19/Wales/PHWC-15DA9C/2020\|EPI_ISL_472757\|2020 | .................... | .................... | .................... | .................... | .................... | .................... | |
| D | 5330 | hCoV-19/Wales/PHWC-15DABA/2020\|EPI_ISL_472758\|2020 | .................... | .................... | .................... | .................... | .................... | .................... | |
| D | 5331 | hCoV-19/Wales/PHWC-15DAF6/2020\|EPI_ISL_472760\|2020 | .................... | .................... | .................... | .................... | .................... | .................... | |
| D | 5332 | hCoV-19/Wales/PHWC-15DB11/2020\|EPI_ISL_472761\|2020 | .................... | .................... | .................... | .................... | NNN................. | .................... | 1 |
| D | 5333 | hCoV-19/Wales/PHWC-15DB6C/2020\|EPI_ISL_472762\|2020 | .................... | .................... | .................... | .................... | .................... | .................... | |
| D | 5334 | hCoV-19/Wales/PHWC-15DB7B/2020\|EPI_ISL_472763\|2020 | .................... | .................... | .................... | .................... | .................... | .................... | |
| D | 5335 | hCoV-19/Wales/PHWC-15DBA8/2020\|EPI_ISL_472764\|2020 | .................... | .................... | .................... | .................... | .................... | .................... | |
| D | 5336 | hCoV-19/Wales/PHWC-15DC2D/2020\|EPI_ISL_472766\|2020 | .................... | .................... | .................... | .................... | .................... | .................... | |
| D | 5337 | hCoV-19/Wales/PHWC-15DC78/2020\|EPI_ISL_472768\|2020 | .................... | .................... | .................... | .................... | .................... | .................... | |
| D | 5338 | hCoV-19/Wales/PHWC-15DC87/2020\|EPI_ISL_472769\|2020 | .................... | .................... | .................... | .................... | .................... | .................... | |
| D | 5339 | hCoV-19/Wales/PHWC-15DCC3/2020\|EPI_ISL_472770\|2020 | .................... | .................... | .................... | .................... | .................... | .................... | |
| D | 5340 | hCoV-19/Wales/PHWC-15DCF0/2020\|EPI_ISL_472771\|2020 | .................... | .A.................. | .................... | .................... | .................... | .................... | |
| D | 5341 | hCoV-19/Wales/PHWC-15DD93/2020\|EPI_ISL_472775\|2020 | .................... | .................... | .................... | .................... | .................... | .................... | |
| D | 5342 | hCoV-19/Wales/PHWC-15DDA2/2020\|EPI_ISL_472776\|2020 | .................... | .................... | .................... | .................... | .................... | .................... | |
| D | 5343 | hCoV-19/Wales/PHWC-15DDB1/2020\|EPI_ISL_472777\|2020 | .................... | .................... | .................... | .................... | .................... | .................... | |
| D | 5344 | hCoV-19/Wales/PHWC-15DE27/2020\|EPI_ISL_472778\|2020 | .................... | .................... | .................... | .................... | .................... | .................... | |
| D | 5345 | hCoV-19/Wales/PHWC-15E0C6/2020\|EPI_ISL_472793\|2020 | .................... | .................... | .................... | .................... | .................... | .................... | |
| D | 5346 | hCoV-19/Wales/PHWC-15E10F/2020\|EPI_ISL_472795\|2020 | .................... | .................... | .................... | .................... | .................... | .................... | |
| D | 5347 | hCoV-19/Wales/PHWC-15E178/2020\|EPI_ISL_472798\|2020 | .................... | .................... | .................... | .................... | .................... | .................... | |
| D | 5348 | hCoV-19/Wales/PHWC-15E187/2020\|EPI_ISL_472799\|2020 | .................... | .................... | .................... | .................... | .................... | .................... | |
| D | 5349 | hCoV-19/USA/VA-DCLS-0424/2020\|EPI_ISL_477640\|2020- | .................... | .................... | .................... | .................... | .................... | .................... | |
| D | 5350 | hCoV-19/England/OXON-B06EF/2020\|EPI_ISL_478902\|202 | .................... | .................... | .................... | .................... | .................... | .................... | |
| D | 5351 | hCoV-19/England/OXON-B06FE/2020\|EPI_ISL_478903\|202 | .................... | .................... | ..A................. | .................... | .................... | .................... | |
| D | 5352 | hCoV-19/England/OXON-B0755/2020\|EPI_ISL_478905\|202 | .................... | .................... | .................... | .................... | .................... | .................... | |
| D | 5353 | hCoV-19/England/OXON-B0773/2020\|EPI_ISL_478906\|202 | .................... | .................... | .................... | .................... | .................... | .................... | |
| D | 5354 | hCoV-19/England/OXON-B07BF/2020\|EPI_ISL_478907\|202 | .................... | .................... | .................... | .................... | .................... | .................... | |
| D | 5355 | hCoV-19/England/OXON-B07CE/2020\|EPI_ISL_478908\|202 | .................... | .................... | .................... | .................... | .................... | .................... | |
| D | 5356 | hCoV-19/England/OXON-B07DD/2020\|EPI_ISL_478909\|202 | .................... | .................... | .................... | .................... | .................... | .................... | |
| D | 5357 | hCoV-19/England/OXON-B07EC/2020\|EPI_ISL_478910\|202 | .................... | .................... | .................... | .................... | .................... | .................... | |
| D | 5358 | hCoV-19/England/OXON-B07FB/2020\|EPI_ISL_478911\|202 | .................... | .................... | .................... | .................... | .................... | .................... | |
| D | 5359 | hCoV-19/England/OXON-B0825/2020\|EPI_ISL_478913\|202 | .................... | .................... | .................... | .................... | .................... | .................... | |
| D | 5360 | hCoV-19/England/OXON-B0843/2020\|EPI_ISL_478914\|202 | .................... | .................... | .................... | .................... | .................... | .................... | |
| D | 5361 | hCoV-19/England/OXON-B08DA/2020\|EPI_ISL_478917\|202 | .................... | .................... | .................... | .................... | .................... | .................... | |
| D | 5362 | hCoV-19/England/OXON-B08E9/2020\|EPI_ISL_478918\|202 | .................... | .................... | .................... | .................... | .................... | .................... | |
| D | 5363 | hCoV-19/England/OXON-B0904/2020\|EPI_ISL_478919\|202 | .................... | .................... | .................... | .................... | .................... | .................... | |
| D | 5364 | hCoV-19/England/OXON-B0913/2020\|EPI_ISL_478920\|202 | .................... | .................... | .................... | .................... | .................... | .................... | |
| D | 5365 | hCoV-19/England/OXON-B0922/2020\|EPI_ISL_478921\|202 | .................... | .................... | .................... | .................... | .................... | .................... | |
| D | 5366 | hCoV-19/England/OXON-B095F/2020\|EPI_ISL_478922\|202 | .................... | .................... | .................... | .................... | .................... | .................... | |
| D | 5367 | hCoV-19/England/OXON-B098C/2020\|EPI_ISL_478923\|202 | .................... | .................... | .................... | .................... | .................... | .................... | |
| D | 5368 | hCoV-19/England/OXON-B099B/2020\|EPI_ISL_478924\|202 | .................... | .................... | .................... | .................... | .................... | .................... | |
| D | 5369 | hCoV-19/England/OXON-B09AA/2020\|EPI_ISL_478925\|202 | .................... | .................... | .................... | .................... | .................... | .................... | |
| D | 5370 | hCoV-19/England/OXON-B09B9/2020\|EPI_ISL_478926\|202 | .................... | .................... | .................... | .................... | .................... | .................... | |
| D | 5371 | hCoV-19/England/OXON-B09C8/2020\|EPI_ISL_478927\|202 | .................... | .................... | .................... | .................... | .................... | .................... | |
| D | 5372 | hCoV-19/England/OXON-B0A01/2020\|EPI_ISL_478928\|202 | .................... | .................... | .................... | .................... | .................... | .................... | |
| D | 5373 | hCoV-19/England/OXON-B0A4D/2020\|EPI_ISL_478929\|202 | .................... | .................... | .................... | .................... | .................... | .................... | |
| D | 5374 | hCoV-19/England/OXON-B0E6F/2020\|EPI_ISL_478970\|202 | .................... | .................... | .................... | .................... | .................... | .................... | |
| D | 5375 | hCoV-19/England/OXON-B0E7E/2020\|EPI_ISL_478971\|202 | .................... | .................... | .................... | .................... | .................... | .................... | |
| D | 5376 | hCoV-19/England/OXON-B0E8D/2020\|EPI_ISL_478972\|202 | .................... | .................... | .................... | .................... | .................... | .................... | |
| D | 5377 | hCoV-19/USA/VA-DCLS-0363/2020\|EPI_ISL_463124\|2020- | .................... | .................... | .................... | .................... | .................... | .................... | |
| D | 5378 | hCoV-19/USA/VA-DCLS-0364/2020\|EPI_ISL_463125\|2020- | .................... | .................... | .................... | .................... | .................... | .................... | |
| D | 5379 | hCoV-19/USA/VA-DCLS-0366/2020\|EPI_ISL_463126\|2020- | .................... | .................... | .................... | .................... | .................... | .................... | |
| D | 5380 | hCoV-19/USA/VA-DCLS-0371/2020\|EPI_ISL_463127\|2020- | .................... | .................... | .................... | .................... | .................... | .................... | |
| D | 5381 | hCoV-19/USA/VA-DCLS-0372/2020\|EPI_ISL_463128\|2020- | .................... | .................... | .................... | .................... | .................... | .................... | |
| D | 5382 | hCoV-19/USA/VA-DCLS-0373/2020\|EPI_ISL_463129\|2020- | .................... | .................... | .................... | .................... | .................... | .................... | |
| D | 5383 | hCoV-19/USA/VA-DCLS-0374/2020\|EPI_ISL_463130\|2020- | .................... | .................... | .................... | .................... | .................... | .................... | |
| D | 5384 | hCoV-19/USA/VA-DCLS-0375/2020\|EPI_ISL_463131\|2020- | .................... | .................... | .................... | .................... | .................... | .................... | |
| D | 5385 | hCoV-19/USA/VA-DCLS-0376/2020\|EPI_ISL_463132\|2020- | .................... | .................... | .................... | .................... | .................... | .................... | |
| D | 5386 | hCoV-19/USA/VA-DCLS-0377/2020\|EPI_ISL_463133\|2020- | .................... | .................... | .................... | .................... | .................... | .................... | |
| D | 5387 | hCoV-19/USA/VA-DCLS-0378/2020\|EPI_ISL_463134\|2020- | .................... | .................... | .................... | .................... | .................... | .................... | |
| D | 5388 | hCoV-19/USA/VA-DCLS-0379/2020\|EPI_ISL_463135\|2020- | .................... | .................... | .................... | .................... | .................... | .................... | |
| D | 5389 | hCoV-19/USA/VA-DCLS-0380/2020\|EPI_ISL_463136\|2020- | .................... | .................... | .................... | .................... | .................... | .................... | |
| D | 5390 | hCoV-19/USA/VA-DCLS-0381/2020\|EPI_ISL_463137\|2020- | .................... | .................... | .................... | .................... | .................... | .................... | |
| D | 5391 | hCoV-19/USA/CT-Yale-199/2020\|EPI_ISL_463138\|2020-0 | .................... | .................... | .................... | .................... | .................... | .................... | |
| D | 5392 | hCoV-19/USA/CT-Yale-203/2020\|EPI_ISL_463141\|2020-0 | .................... | .................... | .................... | .................... | .................... | .................... | |
| D | 5393 | hCoV-19/USA/CT-Yale-215/2020\|EPI_ISL_463151\|2020-0 | .................... | .................... | .................... | .................... | .................... | .................... | |
| D | 5394 | hCoV-19/USA/CT-Yale-221/2020\|EPI_ISL_463156\|2020-0 | .................... | .................... | .................... | .................... | .................... | .................... | |
| D | 5395 | hCoV-19/USA/CT-Yale-222/2020\|EPI_ISL_463157\|2020-0 | .................... | .................... | .................... | .................... | .................... | .................... | |
| D | 5396 | hCoV-19/USA/CT-Yale-223/2020\|EPI_ISL_463158\|2020-0 | .................... | .................... | .................... | .................... | .................... | .................... | |
| D | 5397 | hCoV-19/USA/CT-Yale-230/2020\|EPI_ISL_463164\|2020-0 | .................... | .................... | .................... | .................... | .................... | .................... | |
| D | 5398 | hCoV-19/Canada/BC_23745918/2020\|EPI_ISL_463186\|202 | .................... | .................... | .................... | .................... | .................... | .................... | |
| D | 5399 | hCoV-19/Canada/BC_23815516/2020\|EPI_ISL_463187\|202 | .................... | .................... | .................... | .................... | .................... | .................... | |
| D | 5400 | hCoV-19/Canada/BC_23918026/2020\|EPI_ISL_463189\|202 | .................... | .................... | .................... | .................... | .................... | .................... | |
| D | 5401 | hCoV-19/Canada/BC_24044059/2020\|EPI_ISL_463191\|202 | .................... | .................... | .................... | .................... | .................... | .................... | |
| D | 5402 | hCoV-19/Canada/BC_24150410/2020\|EPI_ISL_463193\|202 | .................... | .................... | .................... | .................... | .................... | .................... | |
| D | 5403 | hCoV-19/Canada/BC_24165067/2020\|EPI_ISL_463194\|202 | .................... | .................... | .................... | .................... | .................... | .................... | |

| | | | | | | | | | |
|---|---|---|---|---|---|---|---|---|---|
| D | 5404 hCoV-19/Canada/BC_24190425/2020\|EPI_ISL_463195\|202 | ···················· | ···················· | ···················· | ···················· | ···················· | ···················· | | |
| D | 5405 hCoV-19/Canada/BC_24192775/2020\|EPI_ISL_463196\|202 | ···················· | ···················· | ···················· | ···················· | ···················· | ···················· | | |
| D | 5406 hCoV-19/Canada/BC_24201275/2020\|EPI_ISL_463198\|202 | ···················· | ···················· | ···················· | ···················· | ···················· | ···················· | | |
| D | 5407 hCoV-19/Canada/BC_24225776/2020\|EPI_ISL_463199\|202 | ···················· | ···················· | ···················· | ···················· | ···················· | ···················· | | |
| D | 5408 hCoV-19/England/201060323/2020\|EPI_ISL_464402\|2020 | ···················· | ···················· | ···················· | ···················· | ···················· | ···················· | | |
| D | 5409 hCoV-19/England/201061000/2020\|EPI_ISL_464403\|2020 | ···················· | ···················· | ···················· | ···················· | ···················· | ···················· | | |
| D | 5410 hCoV-19/England/201061034/2020\|EPI_ISL_464404\|2020 | ···················· | ···················· | ···················· | ···················· | ···················· | ···················· | | |
| D | 5412 hCoV-19/England/201080071/2020\|EPI_ISL_464456\|2020 | ···················· | ···················· | ···················· | ···················· | ···················· | ···················· | | |
| D | 5415 hCoV-19/England/201080076/2020\|EPI_ISL_464459\|2020 | ···················· | ···················· | ···················· | ···················· | ···················· | ···················· | | |
| D | 5416 hCoV-19/England/201080077/2020\|EPI_ISL_464460\|2020 | ···················· | ···················· | ···················· | ···················· | ···················· | ···················· | | |
| D | 5417 hCoV-19/England/201080078/2020\|EPI_ISL_464461\|2020 | ···················· | ···················· | ···················· | ···················· | ··N····N·········N·· | ···················· | 1 | 1 |
| D | 5418 hCoV-19/England/201080080/2020\|EPI_ISL_464463\|2020 | ···················· | ···················· | ···················· | ···················· | ···················· | ···················· | | |
| D | 5419 hCoV-19/England/201080081/2020\|EPI_ISL_464464\|2020 | ···················· | ···················· | ···················· | ···················· | ···················· | ···················· | | |
| D | 5421 hCoV-19/England/201080083/2020\|EPI_ISL_464466\|2020 | ···T················ | ···················· | ···················· | ···················· | ···················· | ···················· | | |
| D | 5422 hCoV-19/England/201080091/2020\|EPI_ISL_464467\|2020 | ···················· | ···················· | ···················· | ···················· | ···················· | ···················· | | |
| D | 5425 hCoV-19/England/201080164/2020\|EPI_ISL_464473\|2020 | ···················· | ···················· | ···················· | ···················· | ···················· | ···················· | | |
| D | 5426 hCoV-19/England/201081129/2020\|EPI_ISL_464475\|2020 | ···················· | ···················· | ···················· | ···················· | ···················· | ···················· | | |
| D | 5428 hCoV-19/England/201081250/2020\|EPI_ISL_464479\|2020 | ···················· | ······C············· | ···················· | ···················· | ···················· | ···················· | | |
| D | 5429 hCoV-19/England/20108125404/2020\|EPI_ISL_464480\|20 | ···················· | ······C············· | ···················· | ···················· | ···················· | ···················· | | |
| D | 5430 hCoV-19/England/20108126004/2020\|EPI_ISL_464481\|20 | ···················· | ···················· | ···················· | ···················· | ···················· | ···················· | | |
| D | 5431 hCoV-19/England/20108127304/2020\|EPI_ISL_464482\|20 | ···················· | ···················· | ···················· | ···················· | ···················· | ···················· | | |
| D | 5432 hCoV-19/England/20108134404/2020\|EPI_ISL_464483\|20 | ···················· | ···················· | ···················· | ···················· | ···················· | ···················· | | |
| D | 5434 hCoV-19/England/20109000104/2020\|EPI_ISL_464488\|20 | ···················· | ···················· | ···················· | ···················· | ···················· | ···················· | | |
| D | 5435 hCoV-19/England/20109000204/2020\|EPI_ISL_464489\|20 | ···················· | ···················· | ···················· | ···················· | ···················· | ···················· | | |
| D | 5436 hCoV-19/England/20109001104/2020\|EPI_ISL_464492\|20 | ···················· | ···················· | ···················· | ···················· | ···················· | ···················· | | |
| D | 5437 hCoV-19/England/20109001704/2020\|EPI_ISL_464497\|20 | ···················· | ···················· | ···················· | ···················· | ···················· | ···················· | | |
| D | 5438 hCoV-19/England/20109001804/2020\|EPI_ISL_464498\|20 | ···················· | ···················· | ···················· | ···················· | ···················· | ···················· | | |
| D | 5440 hCoV-19/England/20122017704/2020\|EPI_ISL_465740\|20 | ···················· | ···················· | ···················· | ···················· | ···················· | ···················· | | |
| D | 5441 hCoV-19/England/20124029204/2020\|EPI_ISL_465788\|20 | ···················· | ···················· | ···················· | ···················· | ···················· | ···················· | | |
| D | 5442 hCoV-19/England/20124097004/2020\|EPI_ISL_465794\|20 | ···················· | ···················· | ···················· | ···················· | ···················· | ···················· | | |
| D | 5443 hCoV-19/England/20126004604/2020\|EPI_ISL_465796\|20 | ···················· | ···················· | ···················· | ···················· | ···················· | ···················· | | |
| D | 5444 hCoV-19/Singapore/323ilmn/2020\|EPI_ISL_469100\|2020 | ···················· | ···················· | ···················· | ···················· | ···················· | ···················· | | |
| D | 5446 hCoV-19/Singapore/347/2020\|EPI_ISL_469102\|2020-05- | ···················· | ···················· | ···················· | ···················· | ···················· | ···················· | | |
| D | 5447 hCoV-19/Singapore/348/2020\|EPI_ISL_469103\|2020-03- | ···················· | ···················· | ···················· | ···················· | ···················· | ···················· | | |
| D | 5448 hCoV-19/Singapore/349/2020\|EPI_ISL_469104\|2020-03- | ···················· | ···················· | ···················· | ···················· | ···················· | ···················· | | |
| D | 5449 hCoV-19/Singapore/350/2020\|EPI_ISL_469105\|2020-04- | ···················· | ···················· | ···················· | ···················· | ···················· | ···················· | | |
| D | 5450 hCoV-19/Singapore/351/2020\|EPI_ISL_469106\|2020-03- | ···················· | ···················· | ···················· | ···················· | ···················· | ···················· | | |
| D | 5451 hCoV-19/Singapore/352/2020\|EPI_ISL_469107\|2020-05- | ···················· | ···················· | ···················· | ···················· | ···················· | ···················· | | |
| D | 5452 hCoV-19/Singapore/401/2020\|EPI_ISL_469150\|2020-04- | ···················· | ···················· | ···················· | ···················· | ···················· | ···················· | | |
| D | 5453 hCoV-19/Singapore/402/2020\|EPI_ISL_469151\|2020-04- | ···················· | ···················· | ···················· | ···················· | ···················· | ···················· | | |
| D | 5454 hCoV-19/Singapore/403/2020\|EPI_ISL_469152\|2020-04- | ···················· | ···················· | ···················· | ···················· | ···················· | ···················· | | |
| D | 5455 hCoV-19/Singapore/404/2020\|EPI_ISL_469153\|2020-04- | ···················· | ···················· | ···················· | ···················· | ···················· | ···················· | | |
| D | 5456 hCoV-19/Singapore/405/2020\|EPI_ISL_469154\|2020-04- | ···················· | ···················· | ···················· | ···················· | ···················· | ···················· | | |
| D | 5457 hCoV-19/Singapore/406/2020\|EPI_ISL_469155\|2020-04- | ···················· | ···················· | ···················· | ···················· | ···················· | ···················· | | |
| D | 5458 hCoV-19/England/CAMB-1AB622/2020\|EPI_ISL_470117\|20 | ···················· | ···················· | ···················· | ···················· | ···················· | ···················· | | |
| D | 5459 hCoV-19/England/CAMB-1AB297/2020\|EPI_ISL_470118\|20 | ···················· | ···················· | ···················· | ···················· | ···················· | ···················· | | |
| D | 5460 hCoV-19/England/CAMB-1AA59F/2020\|EPI_ISL_470119\|20 | ···················· | ···················· | ···················· | ···················· | ···················· | ···················· | | |
| D | 5461 hCoV-19/England/CAMB-1AB5DA/2020\|EPI_ISL_470120\|20 | ···················· | ···················· | ···················· | ···················· | ···················· | ···················· | | |
| D | 5462 hCoV-19/England/CAMB-1AAC30/2020\|EPI_ISL_470121\|20 | ···················· | ···················· | ···················· | ···················· | ···················· | ···················· | | |
| D | 5463 hCoV-19/England/CAMB-1ABBBA/2020\|EPI_ISL_470122\|20 | ···················· | ···················· | ···················· | ···················· | ···················· | ···················· | | |
| D | 5464 hCoV-19/England/CAMB-1AB385/2020\|EPI_ISL_470123\|20 | ···················· | ···················· | ···················· | ···················· | ···················· | ···················· | | |
| D | 5465 hCoV-19/England/CAMB-1AA7A8/2020\|EPI_ISL_470124\|20 | ···················· | ·············A······ | ···················· | ···················· | ···················· | ···················· | | |
| D | 5466 hCoV-19/England/CAMB-1AA9FD/2020\|EPI_ISL_470125\|20 | ···················· | ···················· | ···················· | ···················· | ···················· | ···················· | | |
| D | 5467 hCoV-19/England/CAMB-1AAADC/2020\|EPI_ISL_470127\|20 | ···················· | ···················· | ···················· | ···················· | ···················· | ···················· | | |
| D | 5468 hCoV-19/England/CAMB-1AA5CC/2020\|EPI_ISL_470128\|20 | ···················· | ···················· | ···················· | ···················· | ···················· | ···················· | | |
| D | 5469 hCoV-19/England/CAMB-1AB200/2020\|EPI_ISL_470130\|20 | ···················· | ···················· | ···················· | ···················· | ···················· | ···················· | | |
| D | 5470 hCoV-19/DRC/3947/2020\|EPI_ISL_471400\|2020-04-25 | ···················· | ···················· | ···················· | ···················· | ···················· | ···················· | | |
| D | 5471 hCoV-19/DRC/3949/2020\|EPI_ISL_471401\|2020-04-26 | ···················· | ···················· | ···················· | ···················· | ···················· | ···················· | | |
| D | 5472 hCoV-19/DRC/4012/2020\|EPI_ISL_471402\|2020-04-26 | ···················· | ···················· | ···················· | ···················· | ···················· | ···················· | | |
| D | 5473 hCoV-19/DRC/4020/2020\|EPI_ISL_471403\|2020-04-26 | ···················· | ···················· | ···················· | ···················· | ···················· | ···················· | | |
| D | 5475 hCoV-19/DRC/4060/2020\|EPI_ISL_471405\|2020-04-27 | ···················· | ···················· | ···················· | ···················· | ···················· | ···················· | | |
| D | 5476 hCoV-19/DRC/4075/2020\|EPI_ISL_471406\|2020-04-27 | ···················· | ···················· | ···················· | ···················· | ···················· | ···················· | | |
| D | 5477 hCoV-19/DRC/4084/2020\|EPI_ISL_471407\|2020-04-27 | ···················· | ···················· | ···················· | ···················· | ···················· | ···················· | | |
| D | 5478 hCoV-19/DRC/4132/2020\|EPI_ISL_471408\|2020-04-27 | ···················· | ···················· | ···················· | ···················· | ···················· | ···················· | | |
| D | 5479 hCoV-19/DRC/4141/2020\|EPI_ISL_471409\|2020-04-27 | ···················· | ···················· | ···················· | ···················· | ···················· | ···················· | | |
| D | 5480 hCoV-19/DRC/4145/2020\|EPI_ISL_471410\|2020-04-27 | ···················· | ···················· | ···················· | ···················· | ···················· | ···················· | | |
| D | 5481 hCoV-19/DRC/04186/2020\|EPI_ISL_471411\|2020-04-25 | ···················· | ···················· | ···················· | ···················· | ···················· | ···················· | | |
| D | 5482 hCoV-19/DRC/04187/2020\|EPI_ISL_471412\|2020-04-25 | ···················· | ···················· | ···················· | ···················· | ···················· | ···················· | | |
| D | 5483 hCoV-19/DRC/4194/2020\|EPI_ISL_471414\|2020-04-28 | ···················· | ···················· | ···················· | ···················· | ···················· | ···················· | | |
| D | 5484 hCoV-19/DRC/4250/2020\|EPI_ISL_471415\|2020-04-28 | ···················· | ···················· | ···················· | ···················· | ···················· | ···················· | | |
| D | 5485 hCoV-19/Romania/Buzau-291946/2020\|EPI_ISL_471416\|2 | ···················· | ···················· | ···················· | ···················· | ···················· | ···················· | | |
| D | 5486 hCoV-19/Romania/Buzau-292711/2020\|EPI_ISL_471417\|2 | ···················· | ···················· | ···················· | ···················· | ···················· | ···················· | | |
| D | 5487 hCoV-19/Romania/Buzau-293197/2020\|EPI_ISL_471419\|2 | ···················· | ···················· | ···················· | ···················· | ···················· | ···················· | | |
| D | 5488 hCoV-19/South Korea/KCDC2027/2020\|EPI_ISL_471443\|2 | ···················· | ···················· | ···················· | ···················· | ···················· | ···················· | | |
| D | 5489 hCoV-19/South Korea/KCDC2042/2020\|EPI_ISL_471448\|2 | ···················· | ···················· | ···················· | ···················· | ···················· | ···················· | | |
| D | 5490 hCoV-19/South Korea/KCDC2043/2020\|EPI_ISL_471449\|2 | ···················· | ···················· | ···················· | ···················· | ···················· | ···················· | | |
| D | 5491 hCoV-19/South Korea/KCDC2044/2020\|EPI_ISL_471450\|2 | ···················· | ···················· | ···················· | ···················· | ···················· | ···················· | | |
| D | 5492 hCoV-19/South Korea/KCDC2045/2020\|EPI_ISL_471451\|2 | ···················· | ···················· | ···················· | ···················· | ···················· | ···················· | | |

| | | | | | | | | |
|---|---|---|---|---|---|---|---|---|
| D | 5493 hCoV-19/South Korea/KCDC2046/2020\|EPI_ISL_471452\|2 | ···················· | ···················· | ···················· | ···················· | ···················· | ···················· | |
| D | 5494 hCoV-19/South Korea/KCDC2047/2020\|EPI_ISL_471453\|2 | ···················· | ···················· | ···················· | ···················· | ···················· | ···················· | |
| D | 5495 hCoV-19/South Korea/KCDC2048/2020\|EPI_ISL_471454\|2 | ···················· | ···················· | ···················· | ···················· | ···················· | ···················· | |
| D | 5496 hCoV-19/South Korea/KCDC2049/2020\|EPI_ISL_471455\|2 | ···················· | ···················· | ···················· | ···················· | ···················· | ···················· | |
| D | 5497 hCoV-19/Morocco/HMIMV-Rabat102-03/2020\|EPI_ISL_471 | ···················· | ···················· | ···················· | ···················· | ···················· | ···················· | |
| D | 5498 hCoV-19/Morocco/HMIMV-Rabat1025-04/2020\|EPI_ISL_47 | ···················· | ···················· | ···················· | ···················· | ···················· | ···················· | |
| D | 5499 hCoV-19/Morocco/HMIMV-Rabat1429-04/2020\|EPI_ISL_47 | ···················· | ···················· | ···················· | ···················· | ···················· | ···················· | |
| D | 5500 hCoV-19/Morocco/HMIMV-Rabat1435-04/2020\|EPI_ISL_47 | ···················· | ···················· | ···················· | ···················· | ···················· | ···················· | |
| D | 5501 hCoV-19/Morocco/HMIMV-Rabat1462-04/2020\|EPI_ISL_47 | ···················· | ···················· | ···················· | ···················· | ···················· | ···················· | |
| D | 5502 hCoV-19/Wales/PHWC-15D319/2020\|EPI_ISL_472701\|2020 | ···················· | ·············A······ | ···················· | ···················· | ···················· | ···················· | |
| D | 5503 hCoV-19/Wales/PHWC-15D416/2020\|EPI_ISL_472716\|2020 | ···················· | ···················· | ···················· | ···················· | ···················· | ···················· | |
| D | 5504 hCoV-19/Wales/PHWC-15D4E9/2020\|EPI_ISL_472729\|2020 | ···················· | ···················· | ···················· | ···················· | ···················· | ···················· | |
| D | 5505 hCoV-19/Wales/PHWC-15D531/2020\|EPI_ISL_472734\|2020 | ···················· | ···················· | ···················· | ···················· | ···················· | ···················· | |
| D | 5506 hCoV-19/Wales/PHWC-15D56E/2020\|EPI_ISL_472735\|2020 | ···················· | ···················· | ···················· | ···················· | ···················· | ···················· | |
| D | 5507 hCoV-19/Wales/PHWC-15D601/2020\|EPI_ISL_472737\|2020 | ···················· | ···················A | ···················· | ···················· | ···················· | ···················· | 1 |
| D | 5508 hCoV-19/Wales/PHWC-15D689/2020\|EPI_ISL_472740\|2020 | ···················· | ···················· | ···················· | ···················· | ···················· | ···················· | |
| D | 5509 hCoV-19/Wales/PHWC-15D759/2020\|EPI_ISL_472742\|2020 | ···················· | ···················· | ···················· | ···················· | ···················· | ···················· | |
| D | 5510 hCoV-19/Wales/PHWC-15D874/2020\|EPI_ISL_472746\|2020 | ···················· | ···················· | ···················· | ···················· | ···················· | ···················· | |
| D | 5511 hCoV-19/Wales/PHWC-15D883/2020\|EPI_ISL_472747\|2020 | ···················· | ···················· | ···················· | ···················· | ···················· | ···················· | |
| D | 5512 hCoV-19/Wales/PHWC-15D8CF/2020\|EPI_ISL_472749\|2020 | ···················· | ···················· | ···················· | ···················· | ···················· | ···················· | |
| D | 5513 hCoV-19/Wales/PHWC-15DD2A/2020\|EPI_ISL_472773\|2020 | ···················· | ···················· | ···················· | ···················· | ···················· | ···················· | |
| D | 5514 hCoV-19/Wales/PHWC-15DE72/2020\|EPI_ISL_472779\|2020 | ···················· | ···················· | ···················· | ···················· | ···················· | ···················· | |
| D | 5515 hCoV-19/Wales/PHWC-15DE90/2020\|EPI_ISL_472780\|2020 | ···················· | ···················· | ···················· | ···················· | ···················· | ···················· | |
| D | 5516 hCoV-19/Wales/PHWC-15DEEB/2020\|EPI_ISL_472782\|2020 | ···················· | ···················· | ···················· | ···················· | ···················· | ···················· | |
| D | 5517 hCoV-19/Wales/PHWC-15E011/2020\|EPI_ISL_472786\|2020 | ···················· | ····A··············· | ···················· | ···················· | ···················· | ···················· | |
| D | 5518 hCoV-19/Wales/PHWC-15E05D/2020\|EPI_ISL_472789\|2020 | ···················· | ···················· | ···················· | ···················· | ···················· | ···················· | |
| D | 5519 hCoV-19/Wales/PHWC-15E06C/2020\|EPI_ISL_472790\|2020 | ···················· | ···················· | ···················· | ···················· | ···················· | ···················· | |
| D | 5520 hCoV-19/Wales/PHWC-15E099/2020\|EPI_ISL_472791\|2020 | ···················· | ···················· | ···················· | ···················· | ···················· | ···················· | |
| D | 5522 hCoV-19/Wales/PHWC-160173/2020\|EPI_ISL_472901\|2020 | ···················· | ···················· | ···················· | ···················· | ···················· | ···················· | |
| D | 5523 hCoV-19/Wales/PHWC-160182/2020\|EPI_ISL_472902\|2020 | ···················· | ···················· | ···················· | ···················· | ···················· | ···················· | |
| D | 5524 hCoV-19/Wales/PHWC-1601BF/2020\|EPI_ISL_472904\|2020 | ···················· | ···················· | ···················· | ···················· | ···················· | ···················· | |
| D | 5525 hCoV-19/Wales/PHWC-1601DD/2020\|EPI_ISL_472905\|2020 | ···················· | ···················· | ···················· | ···················· | ···················· | ···················· | |
| D | 5526 hCoV-19/Wales/PHWC-1601EC/2020\|EPI_ISL_472906\|2020 | ···················· | ···················· | ···················· | ···················· | ···················· | ···················· | |
| D | 5527 hCoV-19/Sweden/20-08359/2020\|EPI_ISL_475103\|2020-0 | ···················· | ···················· | ···················· | ···················· | ···················· | ···················· | |
| D | 5528 hCoV-19/Sweden/20-08361/2020\|EPI_ISL_475104\|2020-0 | ···················· | ···················· | ···················· | ···················· | ···················· | ···················· | |
| D | 5529 hCoV-19/Sweden/20-08498/2020\|EPI_ISL_475105\|2020-0 | ···················· | ···················· | ···················· | ···················· | ···················· | ···················· | |
| D | 5530 hCoV-19/Sweden/20-08504/2020\|EPI_ISL_475106\|2020-0 | ···················· | ···················· | ···················· | ···················· | ···················· | ···················· | |
| D | 5531 hCoV-19/Sweden/20-08505/2020\|EPI_ISL_475107\|2020-0 | ···················· | ···················· | ···················· | ···················· | ···················· | ···················· | |
| D | 5532 hCoV-19/Sweden/20-08546/2020\|EPI_ISL_475108\|2020-0 | ···················· | ···················· | ···················· | ···················· | ···················· | ···················· | |
| D | 5533 hCoV-19/Sweden/20-08561/2020\|EPI_ISL_475109\|2020-0 | ···················· | ···················· | ···················· | ···················· | ···················· | ···················· | |
| D | 5534 hCoV-19/Sweden/20-50505/2020\|EPI_ISL_475110\|2020-0 | ···················· | ···················· | ···················· | ···················· | ···················· | ···················· | |
| D | 5535 hCoV-19/Sweden/20-08677/2020\|EPI_ISL_475111\|2020-0 | ···················· | ···················· | ···················· | ···················· | ···················· | ···················· | |
| D | 5536 hCoV-19/Sweden/20-08678/2020\|EPI_ISL_475112\|2020-0 | ···················· | ···················· | ···················· | ···················· | ···················· | ···················· | |
| D | 5537 hCoV-19/Sweden/20-08679/2020\|EPI_ISL_475113\|2020-0 | ···················· | ···················· | ···················· | ···················· | ···················· | ···················· | |
| D | 5538 hCoV-19/Sweden/20-08725/2020\|EPI_ISL_475114\|2020-0 | ···················· | ···················· | ···················· | ···················· | ···················· | ···················· | |
| D | 5539 hCoV-19/Sweden/20-50547/2020\|EPI_ISL_475115\|2020-0 | ···················· | ···················· | ···················· | ···················· | ···················· | ···················· | |
| D | 5540 hCoV-19/Sweden/20-08722/2020\|EPI_ISL_475116\|2020-0 | ···················· | ···················· | ···················· | ···················· | ···················· | ···················· | |
| D | 5541 hCoV-19/Sweden/20-08765/2020\|EPI_ISL_475117\|2020-0 | ···················· | ···················· | ···················· | ···················· | ···················· | ···················· | |
| D | 5542 hCoV-19/Sweden/20-50591/2020\|EPI_ISL_475118\|2020-0 | ···················· | ···················· | ···················· | ···················· | ···················· | ···················· | |
| D | 5543 hCoV-19/Sweden/20-50595/2020\|EPI_ISL_475119\|2020-0 | ···················· | ···················· | ···················· | ···················· | ···················· | ···················· | |
| D | 5544 hCoV-19/Sweden/20-08801/2020\|EPI_ISL_475120\|2020-0 | ···················· | ···················· | ···················· | ···················· | ···················· | ···················· | |
| D | 5545 hCoV-19/Sweden/20-08804/2020\|EPI_ISL_475121\|2020-0 | ···················· | ···················· | ···················· | ···················· | ···················· | ···················· | |
| D | 5546 hCoV-19/Sweden/20-51061/2020\|EPI_ISL_475122\|2020-0 | ···················· | ···················· | ···················· | ···················· | ···················· | ···················· | |
| D | 5547 hCoV-19/Sweden/20-08832/2020\|EPI_ISL_475123\|2020-0 | ···················· | ···················· | ···················· | ···················· | ···················· | ···················· | |
| D | 5548 hCoV-19/Sweden/20-08870/2020\|EPI_ISL_475124\|2020-0 | ···················· | ···················· | ···················· | ···················· | ···················· | ···················· | |
| D | 5549 hCoV-19/Sweden/20-08841/2020\|EPI_ISL_475125\|2020-0 | ···················· | ···················· | ···················· | ···················· | ···················· | ···················· | |
| D | 5550 hCoV-19/Sweden/20-51065/2020\|EPI_ISL_475126\|2020-0 | ···················· | ···················· | ···················· | ···················· | ···················· | ···················· | |
| D | 5551 hCoV-19/Sweden/20-51057/2020\|EPI_ISL_475127\|2020-0 | ···················· | ···················· | ···················· | ···················· | ···················· | ···················· | |
| D | 5552 hCoV-19/Sweden/20-50298/2020\|EPI_ISL_475128\|2020-0 | ···················· | ···················· | ···················· | ···················· | ···················· | ···················· | |
| D | 5553 hCoV-19/Sweden/20-50299/2020\|EPI_ISL_475129\|2020-0 | ···················· | ···················· | ···················· | ···················· | ···················· | ···················· | |
| D | 5554 hCoV-19/Sweden/20-50301/2020\|EPI_ISL_475130\|2020-0 | ···················· | ···················· | ···················· | ···················· | ···················· | ···················· | |
| D | 5555 hCoV-19/Sweden/20-50304/2020\|EPI_ISL_475131\|2020-0 | ···················· | ···················· | ···················· | ···················· | ···················· | ···················· | |
| D | 5556 hCoV-19/Sweden/20-50305/2020\|EPI_ISL_475132\|2020-0 | ···················· | ···················· | ···················· | ···················· | ···················· | ···················· | |
| D | 5557 hCoV-19/Sweden/20-50307/2020\|EPI_ISL_475133\|2020-0 | ···················· | ···················· | ···················· | ···················· | ···················· | ···················· | |
| D | 5558 hCoV-19/Sweden/20-50308/2020\|EPI_ISL_475134\|2020-0 | ···················· | ···················· | ···················· | ···················· | ···················· | ···················· | |
| D | 5559 hCoV-19/Sweden/20-50313/2020\|EPI_ISL_475135\|2020-0 | ···················· | ···················· | ···················· | ···················· | ···················· | ···················· | |
| D | 5560 hCoV-19/Sweden/20-50320/2020\|EPI_ISL_475136\|2020-0 | ···················· | ···················· | ···················· | ···················· | ···················· | ···················· | |
| D | 5561 hCoV-19/Sweden/20-50370/2020\|EPI_ISL_475137\|2020-0 | ···················· | ···················· | ···················· | ···················· | ···················· | ···················· | |
| D | 5562 hCoV-19/Sweden/20-50371/2020\|EPI_ISL_475138\|2020-0 | ···················· | ···················· | ···················· | ···················· | ···················· | ···················· | |
| D | 5563 hCoV-19/Sweden/20-50435/2020\|EPI_ISL_475139\|2020-0 | ···················· | ···················· | ···················· | ···················· | ···················· | ···················· | |
| D | 5564 hCoV-19/Sweden/20-50457/2020\|EPI_ISL_475141\|2020-0 | ···················· | ···················· | ···················· | ···················· | ···················· | ···················· | |
| D | 5565 hCoV-19/Sweden/20-50482/2020\|EPI_ISL_475142\|2020-0 | ···················· | ···················· | ···················· | ···················· | ···················· | ···················· | |
| D | 5566 hCoV-19/Sweden/20-50486/2020\|EPI_ISL_475143\|2020-0 | ···················· | ···················· | ···················· | ···················· | ···················· | ···················· | |
| D | 5567 hCoV-19/Sweden/20-50518/2020\|EPI_ISL_475144\|2020-0 | ···················· | ···················· | ···················· | ···················· | ···················· | ···················· | |
| D | 5568 hCoV-19/Sweden/20-50529/2020\|EPI_ISL_475145\|2020-0 | ···················· | ···················· | ···················· | ···················· | ···················· | ···················· | |
| D | 5569 hCoV-19/Sweden/20-50552/2020\|EPI_ISL_475146\|2020-0 | ···················· | ···················· | ···················· | ···················· | ···················· | ···················· | |
| D | 5570 hCoV-19/Sweden/20-50556/2020\|EPI_ISL_475147\|2020-0 | ···················· | ···················· | ···················· | ···················· | ···················· | ···················· | |
| D | 5571 hCoV-19/Sweden/20-50562/2020\|EPI_ISL_475148\|2020-0 | ···················· | ···················· | ···················· | ···················· | ···················· | ···················· | |

| | | | | | | | | |
|---|---|---|---|---|---|---|---|---|
| D | 5572 | hCoV-19/Sweden/20-50600/2020\|EPI_ISL_475149\|2020-0 | ···················· | ···················· | ···················· | ···················· | ···················· | ···················· |
| D | 5573 | hCoV-19/Sweden/20-50601/2020\|EPI_ISL_475150\|2020-0 | ···················· | ···················· | ···················· | ···················· | ···················· | ···················· |
| D | 5574 | hCoV-19/Sweden/20-50615/2020\|EPI_ISL_475151\|2020-0 | ···················· | ···················· | ···················· | ···················· | ···················· | ···················· |
| D | 5575 | hCoV-19/Sweden/20-50659/2020\|EPI_ISL_475152\|2020-0 | ···················· | ···················· | ···················· | ···················· | ···················· | ···················· |
| D | 5576 | hCoV-19/Sweden/20-50664/2020\|EPI_ISL_475153\|2020-0 | ···················· | ···················· | ···················· | ···················· | ···················· | ···················· |
| D | 5577 | hCoV-19/Sweden/20-50669/2020\|EPI_ISL_475154\|2020-0 | ···················· | ···················· | ···················· | ···················· | ···················· | ···················· |
| D | 5578 | hCoV-19/Sweden/20-50673/2020\|EPI_ISL_475155\|2020-0 | ···················· | ···················· | ···················· | ···················· | ···················· | ···················· |
| D | 5579 | hCoV-19/Sweden/20-08842/2020\|EPI_ISL_475156\|2020-0 | ···················· | ···················· | ···················· | ···················· | ···················· | ···················· |
| D | 5580 | hCoV-19/Sweden/20-50900/2020\|EPI_ISL_475157\|2020-0 | ···················· | ···················· | ···················· | ···················· | ···················· | ···················· |
| D | 5581 | hCoV-19/Sweden/20-50863/2020\|EPI_ISL_475158\|2020-0 | ···················· | ···················· | ···················· | ···················· | ···················· | ···················· |
| D | 5582 | hCoV-19/Sweden/20-50860/2020\|EPI_ISL_475159\|2020-0 | ···················· | ···················· | ···················· | ···················· | ···················· | ···················· |
| D | 5583 | hCoV-19/Sweden/20-50931/2020\|EPI_ISL_475160\|2020-0 | ···················· | ···················· | ···················· | ···················· | ···················· | ···················· |
| D | 5584 | hCoV-19/Sweden/20-50933/2020\|EPI_ISL_475161\|2020-0 | ···················· | ···················· | ···················· | ···················· | ···················· | ···················· |
| D | 5585 | hCoV-19/Sweden/20-50935/2020\|EPI_ISL_475162\|2020-0 | ···················· | ···················· | ···················· | ···················· | ···················· | ···················· |
| D | 5586 | hCoV-19/Sweden/20-50936/2020\|EPI_ISL_475163\|2020-0 | ···················· | ···················· | ···················· | ···················· | ···················· | ···················· |
| D | 5587 | hCoV-19/Sweden/20-50996/2020\|EPI_ISL_475164\|2020-0 | ···················· | ···················· | ···················· | ···················· | ···················· | ···················· |
| D | 5588 | hCoV-19/Bangladesh/BCSIR-NILMRC-104/2020\|EPI_ISL_4 | ···················· | ···················· | ···················· | ···················· | ···················· | ···················· |
| D | 5589 | hCoV-19/Bangladesh/BCSIR-NILMRC-107/2020\|EPI_ISL_4 | ···················· | ···················· | ···················· | ···················· | ···················· | ···················· |
| D | 5590 | hCoV-19/Bangladesh/BCSIR-NILMRC-111/2020\|EPI_ISL_4 | ···················· | ···················· | ···················· | ···················· | ···················· | ···················· |
| D | 5591 | hCoV-19/Bangladesh/BCSIR-NILMRC-117/2020\|EPI_ISL_4 | ········T··········· | ···················· | ···················· | ···················· | ···················· | ···················· |
| D | 5592 | hCoV-19/Bangladesh/BCSIR-NILMRC-018/2020\|EPI_ISL_4 | ···················· | ···················· | ···················· | ···················· | ···················· | ···················· |
| D | 5593 | hCoV-19/Bangladesh/BCSIR-NILMRC-017/2020\|EPI_ISL_4 | ···················· | ···················· | ···················· | ···················· | ···C················ | ···················· |
| D | 5594 | hCoV-19/Bangladesh/BCSIR-NILMRC-102/2020\|EPI_ISL_4 | ···················· | ···················· | ···················· | ···················· | ···················· | ···················· |
| D | 5595 | hCoV-19/Bangladesh/BCSIR-NILMRC-109/2020\|EPI_ISL_4 | ···················· | ···················· | ···················· | ···················· | ···················· | ···················· |
| D | 5596 | hCoV-19/Bangladesh/BCSIR-NILMRC-110/2020\|EPI_ISL_4 | ···················· | ···················· | ···················· | ···················· | ···················· | ···················· |
| D | 5597 | hCoV-19/USA/NE-10749/2020\|EPI_ISL_475176\|2020-05-0 | ···················· | ···················· | ···················· | ···················· | ···················· | ···················· |
| D | 5598 | hCoV-19/USA/NE-10760/2020\|EPI_ISL_475178\|2020-05-0 | ···················· | ···················· | ···················· | ···················· | ···················· | ···················· |
| D | 5599 | hCoV-19/USA/NE-10763/2020\|EPI_ISL_475180\|2020-05-0 | ···················· | ···················· | ···················· | ···················· | ···················· | ···················· |
| D | 5601 | hCoV-19/USA/NE-10833/2020\|EPI_ISL_475185\|2020-05-0 | ···················· | ···················· | ···················· | ···················· | ···················· | ···················· |
| D | 5602 | hCoV-19/USA/NE-10834/2020\|EPI_ISL_475186\|2020-05-0 | ···················· | ···················· | ···················· | ···················· | ···················· | ···················· |
| D | 5603 | hCoV-19/USA/NE-10835/2020\|EPI_ISL_475187\|2020-05-0 | ···················· | ···················· | ···················· | ···················· | ···················· | ···················· |
| D | 5604 | hCoV-19/USA/NE-10837/2020\|EPI_ISL_475188\|2020-05-0 | ···················· | ···················· | ···················· | ···················· | ···················· | ···················· |
| D | 5605 | hCoV-19/USA/NE-10851/2020\|EPI_ISL_475189\|2020-05-0 | ···················· | ···················· | ···················· | ···················· | ···················· | ···················· |
| D | 5606 | hCoV-19/Brazil/PR-L16-CD333/2020\|EPI_ISL_476282\|20 | ···················· | ···················· | ···················· | ···················· | ···················· | ···················· |
| D | 5607 | hCoV-19/Brazil/CE-L16-CD344/2020\|EPI_ISL_476288\|20 | ···················· | ···················· | ···················· | ···················· | ···················· | ···················· |
| D | 5608 | hCoV-19/Brazil/CE-L16-CD346/2020\|EPI_ISL_476289\|20 | ···················· | ···················· | ···················· | ···················· | ···················· | ···················· |
| D | 5609 | hCoV-19/Brazil/SC-L17-CD322/2020\|EPI_ISL_476297\|20 | ···················· | ···················· | ···················· | ···················· | ···················· | ···················· |
| D | 5610 | hCoV-19/Brazil/SP-L7-CD67/2020\|EPI_ISL_476435\|2020 | ···················· | ···················· | ···················· | ···················· | ···················· | ···················· |
| D | 5611 | hCoV-19/Brazil/SP-L7-CD72/2020\|EPI_ISL_476439\|2020 | ···················· | ···················· | ···················· | ···················· | ···················· | ···················· |
| D | 5612 | hCoV-19/Brazil/SP-L7-CD79/2020\|EPI_ISL_476445\|2020 | ···················· | ···················· | ···················· | ···················· | ···················· | ···················· |
| D | 5613 | hCoV-19/Brazil/SP-L7-CD81/2020\|EPI_ISL_476446\|2020 | ···················· | ···················· | ···················· | ···················· | ···················· | ···················· |
| D | 5614 | hCoV-19/Brazil/SP-L9-CD118/2020\|EPI_ISL_476469\|202 | ···················· | ···················· | ···················· | ···················· | ···················· | ···················· |
| D | 5615 | hCoV-19/Brazil/SP-L9-CD91/2020\|EPI_ISL_476490\|2020 | ···················· | ···················· | ···················· | ···················· | ···················· | ···················· |
| D | 5616 | hCoV-19/Senegal/62365/2020\|EPI_ISL_476492\|2020-06- | ···················· | ···················· | ···················· | ···················· | ···················· | ···················· |
| D | 5617 | hCoV-19/Senegal/62368/2020\|EPI_ISL_476493\|2020-06- | ···················· | ···················· | ···················· | ···················· | ···················· | ···················· |
| D | 5618 | hCoV-19/Senegal/62872/2020\|EPI_ISL_476494\|2020-06- | ···················· | ···················· | ···················· | ···················· | ···················· | ···················· |
| D | 5619 | hCoV-19/Senegal/62874/2020\|EPI_ISL_476495\|2020-06- | ···················· | ···················· | ···················· | ···················· | ···················· | ···················· |
| D | 5620 | hCoV-19/Argentina/Heritas_HG001/2020\|EPI_ISL_47649 | ···················· | ···················· | ···················· | ···················· | ···················· | ···················· |
| D | 5621 | hCoV-19/Canada/QC_AA4/2020\|EPI_ISL_476498\|2020-04- | ···················· | ···················· | ···················· | ···················· | ···················· | ···················· |
| D | 5622 | hCoV-19/Canada/QC_AC9/2020\|EPI_ISL_476499\|2020-04- | ···················· | ···················· | ···················· | ···················· | ···················· | ···················· |
| D | 5623 | hCoV-19/USA/WA-UW-2595/2020\|EPI_ISL_477721\|2020-03 | ···················· | ···················· | ···················· | ···················· | ···················· | ···················· |
| D | 5624 | hCoV-19/USA/WA-UW-2601/2020\|EPI_ISL_477723\|2020-03 | ···················· | ···················· | ···················· | ···················· | ···················· | ···················· |
| D | 5625 | hCoV-19/USA/WA-UW-2603/2020\|EPI_ISL_477724\|2020-03 | ···················· | ···················· | ···················· | ···················· | ···················· | ···················· |
| D | 5626 | hCoV-19/England/BIRM-61410/2020\|EPI_ISL_477768\|202 | ···················· | ···················· | ···················· | ···················· | ···················· | ···················· |
| D | 5627 | hCoV-19/England/BIRM-6142F/2020\|EPI_ISL_477769\|202 | ···················· | ···················· | ···················· | ···················· | ···················· | ···················· |
| D | 5628 | hCoV-19/England/BIRM-61498/2020\|EPI_ISL_477774\|202 | ···················· | ···················· | ···················· | ···················· | ···················· | ···················· |
| D | 5629 | hCoV-19/England/BIRM-61568/2020\|EPI_ISL_477776\|202 | ···················· | ···················· | ···················· | ···················· | ···················· | ···················· |
| D | 5630 | hCoV-19/England/BIRM-61595/2020\|EPI_ISL_477779\|202 | ···················· | ···················· | ···················· | ···················· | ···················· | ···················· |
| D | 5631 | hCoV-19/England/CAMB-1B4037/2020\|EPI_ISL_477796\|20 | ···················· | ···················· | ···················· | ···················· | ···················· | ···················· |
| D | 5632 | hCoV-19/England/CAMB-1B4EC4/2020\|EPI_ISL_477798\|20 | ···················· | ···················· | ···················· | ···················· | ···················· | ···················· |
| D | 5633 | hCoV-19/USA/VA-DCLS-0297/2020\|EPI_ISL_463100\|2020- | ···················· | ···················· | ···················· | ···················· | ···················· | ···················· |
| D | 5634 | hCoV-19/USA/VA-DCLS-0298/2020\|EPI_ISL_463101\|2020- | ···················· | ···················· | ···················· | ···················· | ···················· | ···················· |
| D | 5635 | hCoV-19/USA/VA-DCLS-0300/2020\|EPI_ISL_463103\|2020- | ···················· | ···················· | ···················· | ···················· | ···················· | ···················· |
| D | 5636 | hCoV-19/USA/VA-DCLS-0301/2020\|EPI_ISL_463104\|2020- | ···················· | ···················· | ···················· | ···················· | ···················· | ···················· |
| D | 5637 | hCoV-19/USA/VA-DCLS-0302/2020\|EPI_ISL_463105\|2020- | ···················· | ···················· | ···················· | ···················· | ···················· | ···················· |
| D | 5638 | hCoV-19/USA/VA-DCLS-0303/2020\|EPI_ISL_463106\|2020- | ···················· | ···················· | ···················· | ···················· | ···················· | ···················· |
| D | 5639 | hCoV-19/USA/VA-DCLS-0304/2020\|EPI_ISL_463107\|2020- | ···················· | ···················· | ···················· | ···················· | ···················· | ···················· |
| D | 5640 | hCoV-19/USA/VA-DCLS-0305/2020\|EPI_ISL_463108\|2020- | ···················· | ···················· | ···················· | ···················· | ···················· | ···················· |
| D | 5641 | hCoV-19/USA/VA-DCLS-0306/2020\|EPI_ISL_463109\|2020- | ···················· | ···················· | ···················· | ···················· | ···················· | ···················· |
| D | 5642 | hCoV-19/USA/VA-DCLS-0307/2020\|EPI_ISL_463110\|2020- | ···················· | ···················· | ···················· | ···················· | ···················· | ···················· |
| D | 5643 | hCoV-19/USA/VA-DCLS-0308/2020\|EPI_ISL_463111\|2020- | ···················· | ···················· | ···················· | ···················· | ···················· | ···················· |
| D | 5644 | hCoV-19/USA/VA-DCLS-0309/2020\|EPI_ISL_463112\|2020- | ···················· | ···················· | ···················· | ···················· | ···················· | ···················· |
| D | 5645 | hCoV-19/USA/VA-DCLS-0310/2020\|EPI_ISL_463113\|2020- | ···················· | ···················· | ···················· | ···················· | ···················· | ···················· |
| D | 5646 | hCoV-19/USA/VA-DCLS-0347/2020\|EPI_ISL_463114\|2020- | ···················· | ···················· | ···················· | ···················· | ···················· | ···················· |
| D | 5647 | hCoV-19/USA/VA-DCLS-0348/2020\|EPI_ISL_463115\|2020- | ···················· | ···················· | ···················· | ···················· | ···················· | ···················· |
| D | 5648 | hCoV-19/USA/VA-DCLS-0349/2020\|EPI_ISL_463116\|2020- | ···················· | ···················· | ···················· | ···················· | ···················· | ···················· |
| D | 5649 | hCoV-19/USA/VA-DCLS-0350/2020\|EPI_ISL_463117\|2020- | ···················· | ···················· | ···················· | ···················· | ···················· | ···················· |
| D | 5650 | hCoV-19/USA/VA-DCLS-0351/2020\|EPI_ISL_463118\|2020- | ···················· | ···················· | ···················· | ···················· | ···················· | ···················· |

| | | | | | | | | | |
|---|---|---|---|---|---|---|---|---|---|
| D | 5651 | hCoV-19/USA/VA-DCLS-0352/2020\|EPI_ISL_463119\|2020- | ···················· | ···················· | ···················· | ···················· | ···················· | ···················· | |
| D | 5652 | hCoV-19/USA/VA-DCLS-0353/2020\|EPI_ISL_463120\|2020- | ···················· | ···················· | ···················· | ···················· | ···················· | ···················· | |
| D | 5653 | hCoV-19/USA/VA-DCLS-0354/2020\|EPI_ISL_463121\|2020- | ···················· | ···················· | ···················· | ···················· | ···················· | ···················· | |
| D | 5654 | hCoV-19/USA/VA-DCLS-0355/2020\|EPI_ISL_463122\|2020- | ···················· | ···················· | ···················· | ···················· | ···················· | ···················· | |
| D | 5655 | hCoV-19/USA/VA-DCLS-0356/2020\|EPI_ISL_463123\|2020- | ···················· | ···················· | ···················· | ···················· | ···················· | ···················· | |
| D | 5658 | hCoV-19/England/201061300/2020\|EPI_ISL_464409\|2020 | ···················· | ···················· | ···················· | ···················· | ···················· | ···················· | |
| D | 5659 | hCoV-19/England/201061453/2020\|EPI_ISL_464411\|2020 | ···················· | ···················· | ···················· | ···················· | ···················· | ··N······N·········· | |
| D | 5661 | hCoV-19/England/201061457/2020\|EPI_ISL_464415\|2020 | ···················· | ···················· | ···················· | ···················· | ···················· | ···················· | |
| D | 5663 | hCoV-19/England/201080001/2020\|EPI_ISL_464417\|2020 | ···················· | ···················· | ···················· | ···················· | ···················· | ···················· | |
| D | 5666 | hCoV-19/England/201080009/2020\|EPI_ISL_464422\|2020 | ···················· | ···················· | ···················· | ···················· | ··················NN | ···················· | 1 |
| D | 5667 | hCoV-19/England/201080010/2020\|EPI_ISL_464423\|2020 | ···················· | ···················· | ···················· | ···················· | ···················· | ···················· | |
| D | 5668 | hCoV-19/England/201080012/2020\|EPI_ISL_464424\|2020 | ···················· | ···················· | ···················· | ···················· | ···················· | ···················· | |
| D | 5669 | hCoV-19/England/201080013/2020\|EPI_ISL_464425\|2020 | ···················· | ···················· | ···················· | ···················· | ···················· | ···················· | |
| D | 5671 | hCoV-19/England/201080017/2020\|EPI_ISL_464427\|2020 | ···················· | ···················· | ···················· | ···················· | ···················· | ···················· | |
| D | 5675 | hCoV-19/England/201080026/2020\|EPI_ISL_464431\|2020 | ···················· | ···················· | ···················· | ···················· | ···················· | ·N·················· | |
| D | 5676 | hCoV-19/England/201080027/2020\|EPI_ISL_464432\|2020 | ···················· | ···················· | ···················· | ···················· | ···················· | ···················· | |
| D | 5678 | hCoV-19/England/201080029/2020\|EPI_ISL_464434\|2020 | ···················· | ···················· | ···················· | ···················· | ···················· | ···················· | |
| D | 5681 | hCoV-19/England/201080039/2020\|EPI_ISL_464438\|2020 | ···················· | ···················· | ···················· | ···················· | ···················· | ···················· | |
| D | 5684 | hCoV-19/England/201080049/2020\|EPI_ISL_464441\|2020 | ···················· | ···················· | ···················· | ···················· | ···················· | ···················· | |
| D | 5685 | hCoV-19/England/201080050/2020\|EPI_ISL_464442\|2020 | ···················· | ···················· | ···················· | ···················· | ···················· | ···················· | |
| D | 5686 | hCoV-19/England/201080051/2020\|EPI_ISL_464443\|2020 | ···················· | ···················· | ···················· | ···················· | ···················· | ···················· | |
| D | 5687 | hCoV-19/England/201080053/2020\|EPI_ISL_464444\|2020 | ···················· | ···················· | ···················· | ···················· | ···················· | ···················· | |
| D | 5689 | hCoV-19/England/201080056/2020\|EPI_ISL_464447\|2020 | ···················· | ···················· | ···················· | ···················· | ···················· | ···················· | |
| D | 5691 | hCoV-19/England/201080059/2020\|EPI_ISL_464449\|2020 | ···················· | ···················· | ···················· | ···················· | ···················· | ···················· | |
| D | 5692 | hCoV-19/England/201080061/2020\|EPI_ISL_464450\|2020 | ···················· | ···················· | ···················· | ···················· | ···················· | ···················· | |
| D | 5694 | hCoV-19/England/201080064/2020\|EPI_ISL_464452\|2020 | ···················· | ···················· | ···················· | ···················· | ···················· | ···················· | |
| D | 5695 | hCoV-19/Canada/Qc-L00241214/2020\|EPI_ISL_465700\|20 | ···················· | ·A·················· | ···················· | ···················· | ···················· | ···················· | |
| D | 5696 | hCoV-19/Canada/Qc-L00241251/2020\|EPI_ISL_465701\|20 | ···················· | ···················· | ···················· | ···················· | ···················· | ···················· | |
| D | 5697 | hCoV-19/Canada/Qc-L00241319/2020\|EPI_ISL_465702\|20 | ···················· | ···················· | ···················· | ···················· | ···················· | ···················· | |
| D | 5698 | hCoV-19/Canada/Qc-L00241349/2020\|EPI_ISL_465703\|20 | ···················· | ···················· | ···················· | ···················· | ···················· | ···················· | |
| D | 5699 | hCoV-19/England/20186080304/2020\|EPI_ISL_465704\|20 | ···················· | ···················· | ···················· | ···················· | ···················· | ···················· | |
| D | 5700 | hCoV-19/England/20186081804/2020\|EPI_ISL_465706\|20 | ···················· | ···················· | ···················· | ···················· | ···················· | ···················· | |
| D | 5701 | hCoV-19/England/20186084004/2020\|EPI_ISL_465708\|20 | ···················· | ···················· | ···················· | ···················· | ···················· | ···················· | |
| D | 5702 | hCoV-19/England/20186084504/2020\|EPI_ISL_465709\|20 | ···················· | ···················· | ···················· | ···················· | ···················· | ···················· | |
| D | 5703 | hCoV-19/England/20186120904/2020\|EPI_ISL_465710\|20 | ···················· | ···················· | ···················· | ···················· | ···················· | ···················· | |
| D | 5704 | hCoV-19/England/20186133904/2020\|EPI_ISL_465712\|20 | ···················· | ···················· | ···················· | ···················· | ···················· | ···················· | |
| D | 5705 | hCoV-19/England/20186134404/2020\|EPI_ISL_465713\|20 | ···················· | ···················· | ···················· | ···················· | ···················· | ···················· | |
| D | 5706 | hCoV-19/England/20188019604/2020\|EPI_ISL_465714\|20 | ···················· | ···················· | ···················· | ···················· | ···················· | ···················· | |
| D | 5707 | hCoV-19/England/20188028704/2020\|EPI_ISL_465717\|20 | ···················· | ···················· | ···················· | ···················· | ···················· | ···················· | |
| D | 5708 | hCoV-19/England/20188033304/2020\|EPI_ISL_465718\|20 | ···················· | ···················· | ···················· | ···················· | ···················· | ···················· | |
| D | 5710 | hCoV-19/England/20120106104/2020\|EPI_ISL_465728\|20 | ···················· | ···················· | ···················· | ···················· | ···················· | ···················· | |
| D | 5711 | hCoV-19/England/20120107804/2020\|EPI_ISL_465732\|20 | ···················· | ···················· | ···················· | ···················· | ···················· | ···················· | |
| D | 5712 | hCoV-19/England/20122006104/2020\|EPI_ISL_465734\|20 | ···················· | ···················· | ···················· | ···················· | ···················· | ···················· | |
| D | 5716 | hCoV-19/England/20122032304/2020\|EPI_ISL_465754\|20 | ···················· | ···················· | ···················· | ···················· | ···················· | ···················· | |
| D | 5717 | hCoV-19/England/20122054604/2020\|EPI_ISL_465757\|20 | ···················· | ···················· | ···················· | ···················· | ···················· | ···················· | |
| D | 5719 | hCoV-19/England/20122103304/2020\|EPI_ISL_465766\|20 | ···················· | ···················· | ···················· | ···················· | ···················· | ···················· | |
| D | 5720 | hCoV-19/England/20122120604/2020\|EPI_ISL_465767\|20 | ···················· | ···················· | ···················· | ···················· | ···················· | ···················· | |
| D | 5722 | hCoV-19/England/20124000504/2020\|EPI_ISL_465769\|20 | ···················· | ···················· | ···················· | ···················· | ···················· | ···················· | |
| D | 5723 | hCoV-19/England/20124003604/2020\|EPI_ISL_465774\|20 | ···················· | ···················· | ···················· | ···················· | ···················· | ···················· | |
| D | 5725 | hCoV-19/Canada/NL_87/2020\|EPI_ISL_469229\|2020-03-2 | ···················· | ···················· | ···················· | ···················· | ···················· | ···················· | |
| D | 5726 | hCoV-19/Canada/NL_88/2020\|EPI_ISL_469230\|2020-03-2 | ···················· | ···················· | ···················· | ···················· | ···················· | ···················· | |
| D | 5727 | hCoV-19/Canada/NL_89/2020\|EPI_ISL_469231\|2020-03-2 | ···················· | ···················· | ···················· | ···················· | ···················· | ···················· | |
| D | 5728 | hCoV-19/Canada/NL_90/2020\|EPI_ISL_469232\|2020-03-2 | ···················· | ···················· | ···················· | ···················· | ···················· | ···················· | |
| D | 5729 | hCoV-19/Canada/NL_93/2020\|EPI_ISL_469233\|2020-03-2 | ···················· | ···················· | ···················· | ···················· | ···················· | ···················· | |
| D | 5730 | hCoV-19/Canada/NL_95/2020\|EPI_ISL_469234\|2020-03-2 | ···················· | ···················· | ···················· | ···················· | ···················· | ···················· | |
| D | 5731 | hCoV-19/Canada/NL_96/2020\|EPI_ISL_469235\|2020-03-2 | ···················· | ···················· | ···················· | ···················· | ···················· | ···················· | |
| D | 5732 | hCoV-19/Canada/NL_97/2020\|EPI_ISL_469236\|2020-03-2 | ···················· | ···················· | ···················· | ···················· | ···················· | ···················· | |
| D | 5733 | hCoV-19/Canada/NL_98/2020\|EPI_ISL_469237\|2020-03-2 | ···················· | ···················· | ···················· | ···················· | ···················· | ···················· | |
| D | 5734 | hCoV-19/Canada/NL_99/2020\|EPI_ISL_469238\|2020-03-2 | ···················· | ···················· | ···················· | ···················· | ···················· | ···················· | |
| D | 5735 | hCoV-19/Canada/NL_100/2020\|EPI_ISL_469239\|2020-03- | ···················· | ···················· | ···················· | ···················· | ···················· | ···················· | |
| D | 5736 | hCoV-19/Canada/NL_101/2020\|EPI_ISL_469240\|2020-03- | ···················· | ···················· | ···················· | ···················· | ···················· | ···················· | |
| D | 5737 | hCoV-19/Saudi Arabia/4637/2020\|EPI_ISL_469241\|2020 | ···················· | ·A·················· | ···················· | ···················· | ···················· | ···A················ | |
| D | 5738 | hCoV-19/Saudi Arabia/42952/2020\|EPI_ISL_469242\|202 | ···················· | ···················· | ···················· | ···················· | ···················· | ···················· | |
| D | 5739 | hCoV-19/Saudi Arabia/85613/2020\|EPI_ISL_469243\|202 | ···················· | ···················· | ···················· | ···················· | ···················· | ···················· | |
| D | 5740 | hCoV-19/Saudi Arabia/85715/2020\|EPI_ISL_469244\|202 | ···················· | ···················· | ···················· | ···················· | ···················· | ···················· | |
| D | 5741 | hCoV-19/Saudi Arabia/85790/2020\|EPI_ISL_469245\|202 | ···················· | ···················· | ···················· | ···················· | ···················· | ···················· | |
| D | 5742 | hCoV-19/Saudi Arabia/85791/2020\|EPI_ISL_469246\|202 | ···················· | ···················· | ···················· | ···················· | ···················· | ···················· | |
| D | 5743 | hCoV-19/Saudi Arabia/86267/2020\|EPI_ISL_469247\|202 | ···················· | ···················· | ···················· | ···················· | ···················· | ···················· | |
| D | 5744 | hCoV-19/Saudi Arabia/86327/2020\|EPI_ISL_469248\|202 | ···················· | ···················· | ···················· | ···················· | ···················· | ···················· | |
| D | 5745 | hCoV-19/Saudi Arabia/86650/2020\|EPI_ISL_469249\|202 | ···················· | ···················· | ···················· | ···················· | ···················· | ···················· | |
| D | 5746 | hCoV-19/Saudi Arabia/832279/2020\|EPI_ISL_469250\|20 | ···················· | ···················· | ···················· | ···················· | ···················· | ···················· | |
| D | 5747 | hCoV-19/Saudi Arabia/85790C/2020\|EPI_ISL_469251\|20 | ···················· | ···················· | ···················· | ···················· | ···················· | ···················· | |
| D | 5748 | hCoV-19/Saudi Arabia/85791C/2020\|EPI_ISL_469252\|20 | ···················· | ···················· | ···················· | ···················· | ···················· | ···················· | |
| D | 5749 | hCoV-19/Shanghai/CH-02/2020\|EPI_ISL_469253\|2020-02 | ···················· | ···················· | ···················· | ···················· | ···················· | ···················· | |
| D | 5750 | hCoV-19/Beijing/IVDC-01-06/2020\|EPI_ISL_469254\|202 | ···················· | ···················· | ···················· | ···················· | ···················· | ···················· | |
| D | 5751 | hCoV-19/Beijing/IVDC-02-06/2020\|EPI_ISL_469255\|202 | ···················· | ···················· | ···················· | ···················· | ···················· | ···················· | |
| D | 5752 | hCoV-19/env/Beijing/IVDC-03-06/2020\|EPI_ISL_469256 | ···················· | ···················· | ···················· | ···················· | ···················· | ···················· | |
| D | 5753 | hCoV-19/Japan/Donner32/2020\|EPI_ISL_469290\|2020-05 | ···················· | ···················· | ···················· | ···················· | ···················· | ···················· | |

| | | | | | | | |
|---|---|---|---|---|---|---|---|
| D | 5754 hCoV-19/Japan/Donner33/2020\|EPI_ISL_469291\|2020-05 | .................... | .................... | ..................... | .................... | .................... | .................... |
| D | 5755 hCoV-19/Japan/Donner34/2020\|EPI_ISL_469292\|2020-05 | .................... | .................... | ..................... | .................... | .................... | .................... |
| D | 5756 hCoV-19/Japan/Donner35/2020\|EPI_ISL_469293\|2020-04 | .................... | .................... | ..................... | .................... | .................... | .................... |
| D | 5757 hCoV-19/Japan/Donner38/2020\|EPI_ISL_469296\|2020-04 | .................... | .................... | ..................... | .................... | .................... | .................... |
| D | 5758 hCoV-19/Bangladesh/BCSIR-NILMRC-096/2020\|EPI_ISL_4 | .................... | .................... | ..................... | .................... | .................... | .................... |
| D | 5759 hCoV-19/Bangladesh/BCSIR-NILMRC-100/2020\|EPI_ISL_4 | .................... | .................... | ..................... | .................... | .................... | .................... |
| D | 5760 hCoV-19/England/CAMB-1AA7D5/2020\|EPI_ISL_470212\|20 | .................... | .................... | ..................... | .................... | .................... | .................... |
| D | 5761 hCoV-19/England/CAMB-1AB798/2020\|EPI_ISL_470214\|20 | .................... | .................... | ..................... | .................... | .................... | .................... |
| D | 5762 hCoV-19/England/CAMB-1AA68D/2020\|EPI_ISL_470215\|20 | .................... | .................... | ..................... | .................... | .................... | .................... |
| D | 5763 hCoV-19/England/CAMB-1AADC4/2020\|EPI_ISL_470217\|20 | .................... | .................... | ..................... | .................... | .................... | .................... |
| D | 5764 hCoV-19/England/CAMB-1AAB33/2020\|EPI_ISL_470218\|20 | .................... | .................... | ..................... | .................... | .................... | .................... |
| D | 5765 hCoV-19/England/CAMB-1ABDA5/2020\|EPI_ISL_470219\|20 | .................... | .................... | ..................... | .................... | .................... | .................... |
| D | 5766 hCoV-19/England/CAMB-1AA77B/2020\|EPI_ISL_470221\|20 | .................... | .................... | ..................... | .................... | .................... | .................... |
| D | 5767 hCoV-19/England/CAMB-1AB15E/2020\|EPI_ISL_470223\|20 | .................... | .................... | ..................... | .................... | .................... | .................... |
| D | 5768 hCoV-19/England/CAMB-1ABA71/2020\|EPI_ISL_470225\|20 | .................... | .................... | ..................... | .................... | .................... | .................... |
| D | 5769 hCoV-19/England/CAMB-1ABD87/2020\|EPI_ISL_470227\|20 | .................... | .................... | ..................... | .................... | .................... | .................... |
| D | 5770 hCoV-19/England/CAMB-1AB91A/2020\|EPI_ISL_470252\|20 | .................... | .................... | ..................... | .................... | .................... | .................... |
| D | 5771 hCoV-19/England/CAMB-1AA5F9/2020\|EPI_ISL_470254\|20 | .................... | .................... | ..................... | .................... | .................... | .................... |
| D | 5772 hCoV-19/England/CAMB-1AB31C/2020\|EPI_ISL_470255\|20 | .................... | .................... | ..................... | .................... | .................... | .................... |
| D | 5773 hCoV-19/England/CAMB-1AB30D/2020\|EPI_ISL_470256\|20 | .................... | .................... | ..................... | .................... | .................... | .................... |
| D | 5774 hCoV-19/England/CAMB-1ABD78/2020\|EPI_ISL_470258\|20 | .................... | .................... | ..................... | .................... | .................... | .................... |
| D | 5775 hCoV-19/England/CAMB-1AB886/2020\|EPI_ISL_470259\|20 | .................... | .................... | ..................... | .................... | .................... | .................... |
| D | 5776 hCoV-19/England/CAMB-1AAE1C/2020\|EPI_ISL_470260\|20 | .................... | .................... | ..................... | .................... | .................... | .................... |
| D | 5777 hCoV-19/England/CAMB-1ABC6C/2020\|EPI_ISL_470261\|20 | .................... | .................... | ..................... | .................... | .................... | .................... |
| D | 5778 hCoV-19/England/CAMB-1AB1F4/2020\|EPI_ISL_470265\|20 | .................... | .................... | ..................... | .................... | .................... | .................... |
| D | 5779 hCoV-19/England/CAMB-1AB33A/2020\|EPI_ISL_470267\|20 | .................... | .................... | ..................... | .................... | .................... | .................... |
| D | 5780 hCoV-19/England/CAMB-1AB9A1/2020\|EPI_ISL_470268\|20 | .................... | .................... | ..................... | .................... | .................... | .................... |
| D | 5781 hCoV-19/England/CAMB-1ABE2A/2020\|EPI_ISL_470270\|20 | .................... | .................... | ..................... | .................... | .................... | .................... |
| D | 5782 hCoV-19/England/CAMB-1ABC3F/2020\|EPI_ISL_470271\|20 | .................... | .................... | ..................... | .................... | .................... | .................... |
| D | 5783 hCoV-19/England/CAMB-1AB112/2020\|EPI_ISL_470273\|20 | .................... | .................... | ..................... | .................... | .................... | .................... |
| D | 5784 hCoV-19/England/CAMB-1AB7B6/2020\|EPI_ISL_470274\|20 | .................... | .................... | ..................... | .................... | .................... | .................... |
| D | 5785 hCoV-19/England/CAMB-1AB6D7/2020\|EPI_ISL_470275\|20 | .................... | .................... | ..................... | .................... | .................... | .................... |
| D | 5786 hCoV-19/England/CAMB-1AB23D/2020\|EPI_ISL_470276\|20 | .................... | .................... | ..................... | .................... | .................... | .................... |
| D | 5787 hCoV-19/England/CAMB-1AA6D8/2020\|EPI_ISL_470277\|20 | .................... | .................... | ..................... | .................... | .................... | .................... |
| D | 5788 hCoV-19/England/CAMB-1AAC03/2020\|EPI_ISL_470278\|20 | .................... | .................... | ..................... | .................... | .................... | .................... |
| D | 5789 hCoV-19/England/CAMB-1AA614/2020\|EPI_ISL_470279\|20 | .................... | .................... | ..................... | .................... | .................... | .................... |
| D | 5790 hCoV-19/England/CAMB-1AAD3D/2020\|EPI_ISL_470280\|20 | .................... | .................... | ..................... | .................... | .................... | .................... |
| D | 5791 hCoV-19/England/CAMB-1AB8C2/2020\|EPI_ISL_470281\|20 | .................... | .................... | ..................... | .................... | .................... | .................... |
| D | 5792 hCoV-19/England/CAMB-1AAD79/2020\|EPI_ISL_470282\|20 | .................... | .................... | ..................... | .................... | .................... | .................... |
| D | 5793 hCoV-19/England/CAMB-1AAA09/2020\|EPI_ISL_470283\|20 | .................... | .................... | ..................... | .................... | .................... | .................... |
| D | 5794 hCoV-19/England/CAMB-1ABB6F/2020\|EPI_ISL_470284\|20 | .................... | .................... | ..................... | .................... | .................... | .................... |
| D | 5795 hCoV-19/England/CAMB-1ABCD5/2020\|EPI_ISL_470285\|20 | .................... | .................... | ..................... | .................... | .................... | .................... |
| D | 5796 hCoV-19/England/CAMB-1AA7E4/2020\|EPI_ISL_470286\|20 | .................... | .................... | ..................... | .................... | .................... | .................... |
| D | 5797 hCoV-19/England/CAMB-1AACB8/2020\|EPI_ISL_470289\|20 | .................... | .................... | ..................... | .................... | .................... | .................... |
| D | 5798 hCoV-19/England/CAMB-1AB4EC/2020\|EPI_ISL_470290\|20 | .................... | .................... | ..................... | .................... | .................... | .................... |
| D | 5799 hCoV-19/England/CAMB-1ABCC6/2020\|EPI_ISL_470291\|20 | .................... | .................... | ..................... | .................... | .................... | .................... |
| D | 5800 hCoV-19/England/CAMB-1AAD4C/2020\|EPI_ISL_470292\|20 | .................... | .................... | ..................... | .................... | .................... | .................... |
| D | 5801 hCoV-19/England/CAMB-1AA75D/2020\|EPI_ISL_470293\|20 | .................... | .................... | ..................... | .................... | .................... | .................... |
| D | 5802 hCoV-19/England/CAMB-1AB2B5/2020\|EPI_ISL_470297\|20 | .................... | .................... | ..................... | .................... | .................... | .................... |
| D | 5803 hCoV-19/England/CAMB-1AAA81/2020\|EPI_ISL_470298\|20 | .................... | .................... | ..................... | .................... | .................... | .................... |
| D | 5804 hCoV-19/England/CAMB-1AAA45/2020\|EPI_ISL_470299\|20 | .................... | .................... | ..................... | .................... | .................... | .................... |
| D | 5805 hCoV-19/Czech Republic/NRL_2307/2020\|EPI_ISL_47154 | .................... | .................... | ..................... | .................... | .................... | .................... |
| D | 5806 hCoV-19/Brazil/SP-126/2020\|EPI_ISL_471541\|2020-04- | .................... | .................... | ..................... | .................... | .................... | .................... |
| D | 5807 hCoV-19/Brazil/SP-127/2020\|EPI_ISL_471542\|2020-04- | .................... | .................... | ..................... | .................... | .................... | .................... |
| D | 5808 hCoV-19/Brazil/SP-131/2020\|EPI_ISL_471543\|2020-04- | .................... | .................... | ..................... | .................... | .................... | .................... |
| D | 5809 hCoV-19/Czech Republic/NRL_2312/2020\|EPI_ISL_47154 | .................... | .................... | ..................... | .................... | .................... | .................... |
| D | 5810 hCoV-19/Brazil/SP-601/2020\|EPI_ISL_471545\|2020-03- | .................... | .................... | ..................... | .................... | .................... | .................... |
| D | 5811 hCoV-19/Brazil/SP-537/2020\|EPI_ISL_471546\|2020-03- | .................... | .................... | ..................... | .................... | .................... | .................... |
| D | 5812 hCoV-19/Czech Republic/NRL_2554/2020\|EPI_ISL_47154 | .................... | .................... | ..................... | .................... | .................... | .................... |
| D | 5813 hCoV-19/Brazil/SP-606/2020\|EPI_ISL_471548\|2020-04- | .................... | .................... | ..................... | .................... | .................... | .................... |
| D | 5814 hCoV-19/Brazil/SP-607/2020\|EPI_ISL_471549\|2020-04- | .................... | .................... | ..................... | .................... | .................... | .................... |
| D | 5815 hCoV-19/Czech Republic/NRL_5240/2020\|EPI_ISL_47155 | .................... | .................... | ..................... | .................... | .................... | .................... |
| D | 5816 hCoV-19/Brazil/SP-545/2020\|EPI_ISL_471551\|2020-04- | .................... | .................... | ..................... | .................... | .................... | .................... |
| D | 5817 hCoV-19/Brazil/SP-549/2020\|EPI_ISL_471552\|2020-04- | .................... | .................... | ..................... | .................... | .................... | .................... |
| D | 5818 hCoV-19/Czech Republic/NRL_5186/2020\|EPI_ISL_47155 | .................... | .................... | ..................... | .................... | .................... | .................... |
| D | 5819 hCoV-19/Brazil/SP-551/2020\|EPI_ISL_471554\|2020-03- | .................... | .................... | ..................... | .................... | .................... | .................... |
| D | 5820 hCoV-19/Czech Republic/NRL_5184/2020\|EPI_ISL_47155 | .................... | .................... | ..................... | .................... | .................... | .................... |
| D | 5821 hCoV-19/Brazil/SP-512/2020\|EPI_ISL_471556\|2020-04- | .................... | .................... | ...........T......... | .................... | .................... | .................... |
| D | 5822 hCoV-19/India/TN-CCMB-C20/2020\|EPI_ISL_471583\|2020 | .................... | .................... | ..................... | .................... | .................... | .................... |
| D | 5823 hCoV-19/India/TN-CCMB-C21/2020\|EPI_ISL_471584\|2020 | .................... | .................... | ..................... | .................... | .................... | .................... |
| D | 5824 hCoV-19/India/TG-CCMB-J812-P2/2020\|EPI_ISL_471585\| | .................... | .................... | ..................... | .................... | .................... | .................... |
| D | 5825 hCoV-19/India/TG-CCMB-L040/2020\|EPI_ISL_471586\|202 | .................... | .................... | ..................... | .................... | .................... | .................... |
| D | 5826 hCoV-19/India/TG-CCMB-L040-P3/2020\|EPI_ISL_471587\| | .................... | .................... | ..................... | .................... | .................... | .................... |
| D | 5827 hCoV-19/India/TG-CCMB-L1208/2020\|EPI_ISL_471588\|20 | .................... | .A.................. | ..................... | .................... | .................... | .................... |
| D | 5828 hCoV-19/India/TG-CCMB-L1276/2020\|EPI_ISL_471589\|20 | .................... | .A.................. | ..................... | .................... | .................... | .................... |
| D | 5829 hCoV-19/India/TG-CCMB-L1451/2020\|EPI_ISL_471590\|20 | .................... | .................... | ..................... | .................... | .................... | .................... |
| D | 5830 hCoV-19/India/TG-CCMB-L1452/2020\|EPI_ISL_471591\|20 | .................... | .................... | ..................... | .................... | .................... | .................... |
| D | 5831 hCoV-19/India/TG-CCMB-L1453/2020\|EPI_ISL_471592\|20 | .................... | .................... | ..................... | .................... | .................... | .................... |

| | | | | | | | |
|---|---|---|---|---|---|---|---|
| D | 5832 hCoV-19/India/TG-CCMB-L1454/2020\|EPI_ISL_471593\|20 | .................... | .................... | .................... | .................... | .................... | .................... |
| D | 5833 hCoV-19/India/TG-CCMB-L1455/2020\|EPI_ISL_471594\|20 | .................... | .................... | .................... | .................... | .................... | .................... |
| D | 5834 hCoV-19/India/TG-CCMB-L1456/2020\|EPI_ISL_471595\|20 | .................... | .................... | .................... | .................... | .................... | .................... |
| D | 5835 hCoV-19/India/TG-CCMB-L1459/2020\|EPI_ISL_471596\|20 | .................... | .................... | .................... | .................... | .................... | .................... |
| D | 5836 hCoV-19/India/TG-CCMB-L1460/2020\|EPI_ISL_471597\|20 | .................... | .................... | .................... | .................... | .................... | .................... |
| D | 5837 hCoV-19/India/TG-CCMB-L1465/2020\|EPI_ISL_471598\|20 | .................... | .................... | .................... | .................... | .................... | .................... |
| D | 5838 hCoV-19/India/TG-CCMB-L1466/2020\|EPI_ISL_471599\|20 | .................... | .................... | .................... | .................... | .................... | .................... |
| D | 5839 hCoV-19/Wales/PHWC-15FEF8/2020\|EPI_ISL_472870\|2020 | .................... | .................... | .................... | .................... | .................... | .................... |
| D | 5840 hCoV-19/Wales/PHWC-15FF40/2020\|EPI_ISL_472873\|2020 | .................... | .................... | .................... | .................... | .................... | .................... |
| D | 5841 hCoV-19/Wales/PHWC-15FF5F/2020\|EPI_ISL_472874\|2020 | .................... | .................... | .................... | .................... | .................... | .................... |
| D | 5842 hCoV-19/Wales/PHWC-15FF6E/2020\|EPI_ISL_472875\|2020 | .................... | .................... | .................... | .................... | .................... | .................... |
| D | 5843 hCoV-19/Wales/PHWC-15FFB9/2020\|EPI_ISL_472878\|2020 | .................... | .................... | .................... | .................... | .................... | .................... |
| D | 5844 hCoV-19/Wales/PHWC-15FFD7/2020\|EPI_ISL_472879\|2020 | .................... | .................... | .................... | .................... | .................... | .................... |
| D | 5845 hCoV-19/Wales/PHWC-15FFE6/2020\|EPI_ISL_472880\|2020 | .................... | .................... | .................... | .................... | .................... | .................... |
| D | 5846 hCoV-19/Wales/PHWC-16000D/2020\|EPI_ISL_472882\|2020 | .................... | .................... | .................... | .................... | .................... | .................... |
| D | 5847 hCoV-19/Wales/PHWC-16001C/2020\|EPI_ISL_472883\|2020 | .................... | .................... | .................... | .................... | .................... | .................... |
| D | 5848 hCoV-19/Wales/PHWC-16002B/2020\|EPI_ISL_472884\|2020 | .................... | .................... | .................... | .................... | .................... | .................... |
| D | 5849 hCoV-19/Wales/PHWC-160049/2020\|EPI_ISL_472886\|2020 | .................... | .................... | .................... | .................... | .................... | .................... |
| D | 5850 hCoV-19/Wales/PHWC-160076/2020\|EPI_ISL_472889\|2020 | .................... | .................... | .................... | .................... | .................... | .................... |
| D | 5851 hCoV-19/Sweden/20-08461/2020\|EPI_ISL_475100\|2020-0 | .................... | .................... | .................... | .................... | .................... | .................... |
| D | 5852 hCoV-19/Sweden/20-08357/2020\|EPI_ISL_475101\|2020-0 | .................... | .................... | .................... | .................... | .................... | .................... |
| D | 5853 hCoV-19/Sweden/20-08358/2020\|EPI_ISL_475102\|2020-0 | .................... | .................... | .................... | .................... | .................... | .................... |
| D | 5854 hCoV-19/USA/WA-UW-2056/2020\|EPI_ISL_477700\|2020-03 | .................... | .................... | .................... | .................... | .................... | .................... |
| D | 5855 hCoV-19/USA/WA-UW-2059/2020\|EPI_ISL_477701\|2020-03 | .................... | .................... | .................... | .................... | .................... | .................... |
| D | 5856 hCoV-19/USA/WA-UW-2062/2020\|EPI_ISL_477702\|2020-03 | .................... | .................... | .................... | .................... | .................... | .................... |
| D | 5857 hCoV-19/USA/WA-UW-2066/2020\|EPI_ISL_477703\|2020-03 | .................... | .................... | .................... | .................... | .................... | .................... |
| D | 5858 hCoV-19/USA/WA-UW-2076/2020\|EPI_ISL_477704\|2020-03 | .................... | .................... | .................... | .................... | .................... | .................... |
| D | 5859 hCoV-19/USA/WA-UW-2268/2020\|EPI_ISL_477705\|2020-03 | .................... | .................... | .................... | .................... | .................... | .................... |
| D | 5860 hCoV-19/USA/WA-UW-2270/2020\|EPI_ISL_477706\|2020-03 | .................... | .................... | .................... | .................... | .................... | .................... |
| D | 5861 hCoV-19/USA/WA-UW-2276/2020\|EPI_ISL_477707\|2020-03 | .................... | .................... | .................... | .................... | .................... | .................... |
| D | 5862 hCoV-19/USA/WA-UW-2280/2020\|EPI_ISL_477708\|2020-03 | .................... | .................... | .................... | .................... | .................... | .................... |
| D | 5863 hCoV-19/USA/WA-UW-2281/2020\|EPI_ISL_477709\|2020-03 | .................... | .................... | .................... | .................... | .................... | .................... |
| D | 5864 hCoV-19/USA/WA-UW-2293/2020\|EPI_ISL_477710\|2020-03 | .................... | .................... | .................... | .................... | .................... | .................... |
| D | 5865 hCoV-19/USA/WA-UW-2317/2020\|EPI_ISL_477711\|2020-03 | .................... | .................... | .................... | .................... | .................... | .................... |
| D | 5866 hCoV-19/USA/WA-UW-2320/2020\|EPI_ISL_477712\|2020-03 | .................... | .................... | .................... | .................... | .................... | .................... |
| D | 5867 hCoV-19/USA/WA-UW-2370/2020\|EPI_ISL_477713\|2020-03 | .................... | .................... | .................... | .................... | .................... | .................... |
| D | 5868 hCoV-19/USA/WA-UW-2374/2020\|EPI_ISL_477714\|2020-03 | .................... | .................... | .................... | .................... | .................... | .................... |
| D | 5869 hCoV-19/USA/WA-UW-2376/2020\|EPI_ISL_477715\|2020-03 | .................... | .................... | .................... | .................... | .................... | .................... |
| D | 5870 hCoV-19/USA/WA-UW-2386/2020\|EPI_ISL_477716\|2020-03 | .................... | .................... | .................... | .................... | .................... | .................... |
| D | 5871 hCoV-19/USA/WA-UW-2521/2020\|EPI_ISL_477717\|2020-03 | .................... | .................... | .................... | .................... | .................... | .................... |
| D | 5872 hCoV-19/USA/WA-UW-2541/2020\|EPI_ISL_477718\|2020-03 | .................... | .................... | .................... | .................... | .................... | .................... |
| D | 5873 hCoV-19/USA/WA-UW-2548/2020\|EPI_ISL_477719\|2020-03 | .................... | .................... | .................... | .................... | .................... | .................... |
| D | 5874 hCoV-19/USA/WA-UW-2557/2020\|EPI_ISL_477720\|2020-03 | .................... | .................... | .................... | .................... | .................... | .................... |
| D | 5875 hCoV-19/England/BIRM-60596/2020\|EPI_ISL_477730\|202 | .................... | .................... | .................... | .................... | .................... | .................... |
| D | 5876 hCoV-19/England/BIRM-605E1/2020\|EPI_ISL_477735\|202 | .................... | .................... | .................... | .................... | .................... | .................... |
| D | 5877 hCoV-19/England/BIRM-60693/2020\|EPI_ISL_477741\|202 | .................... | .................... | .................... | .................... | .................... | .................... |
| D | 5878 hCoV-19/England/BIRM-606A2/2020\|EPI_ISL_477742\|202 | .................... | .................... | .................... | .................... | .................... | .................... |
| D | 5879 hCoV-19/England/BIRM-61094/2020\|EPI_ISL_477744\|202 | .................... | .................... | .................... | .................... | .................... | .................... |
| D | 5880 hCoV-19/England/BIRM-610B2/2020\|EPI_ISL_477746\|202 | .................... | .................... | .................... | .................... | .................... | .................... |
| D | 5881 hCoV-19/England/BIRM-610C1/2020\|EPI_ISL_477747\|202 | .................... | .................... | .................... | .................... | .................... | .................... |
| D | 5882 hCoV-19/England/BIRM-610FE/2020\|EPI_ISL_477749\|202 | .................... | .................... | .................... | .................... | .................... | .................... |
| D | 5883 hCoV-19/England/BIRM-61119/2020\|EPI_ISL_477751\|202 | .................... | .................... | .................... | .................... | .................... | .................... |
| D | 5884 hCoV-19/England/BIRM-61128/2020\|EPI_ISL_477752\|202 | .................... | .................... | .................... | .................... | .................... | .................... |
| D | 5885 hCoV-19/England/BIRM-612DA/2020\|EPI_ISL_477756\|202 | .................... | .................... | .................... | .................... | .................... | .................... |
| D | 5886 hCoV-19/England/BIRM-612F8/2020\|EPI_ISL_477758\|202 | .................... | .................... | .................... | .................... | .................... | .................... |
| D | 5887 hCoV-19/England/BIRM-61331/2020\|EPI_ISL_477762\|202 | .................... | .................... | .................... | .................... | .................... | .................... |
| D | 5888 hCoV-19/England/BIRM-61340/2020\|EPI_ISL_477763\|202 | .................... | .................... | .................... | .................... | .................... | .................... |
| D | 5889 hCoV-19/USA/VA-DCLS-0288/2020\|EPI_ISL_463094\|2020- | .................... | .................... | .................... | .................... | .................... | .................... |
| D | 5890 hCoV-19/USA/VA-DCLS-0289/2020\|EPI_ISL_463095\|2020- | .................... | .................... | .................... | .................... | .................... | .................... |
| D | 5891 hCoV-19/USA/VA-DCLS-0292/2020\|EPI_ISL_463096\|2020- | .................... | .................... | .................... | .................... | .................... | .................... |
| D | 5892 hCoV-19/USA/VA-DCLS-0295/2020\|EPI_ISL_463098\|2020- | .................... | .................... | .................... | .................... | .................... | .................... |
| D | 5893 hCoV-19/USA/VA-DCLS-0296/2020\|EPI_ISL_463099\|2020- | .................... | .................... | .................... | .................... | .................... | .................... |
| D | 5894 hCoV-19/South Africa/Tygerberg_01/2020\|EPI_ISL_464 | .................... | .................... | .................... | .................... | .................... | .................... |
| D | 5895 hCoV-19/South Africa/Tygerberg_02/2020\|EPI_ISL_464 | .................... | .A.................. | .................... | .................... | .................... | .................... |
| D | 5896 hCoV-19/South Africa/Tygerberg_03/2020\|EPI_ISL_464 | .................... | .A.................. | .................... | .................... | .................... | .................... |
| D | 5897 hCoV-19/South Africa/Tygerberg_07/2020\|EPI_ISL_464 | .................... | .................... | .................... | .................... | .................... | .................... |
| D | 5898 hCoV-19/South Africa/Tygerberg_08/2020\|EPI_ISL_464 | .................... | .................... | .................... | .................... | .................... | .................... |
| D | 5899 hCoV-19/South Africa/Tygerberg_10/2020\|EPI_ISL_464 | .................... | .................... | .................... | .................... | .................... | .................... |
| D | 5900 hCoV-19/South Africa/Tygerberg_12/2020\|EPI_ISL_464 | .................... | .................... | .................... | .................... | .................... | .................... |
| D | 5901 hCoV-19/South Africa/Tygerberg_15/2020\|EPI_ISL_464 | .................... | .................... | .................... | .................... | .................... | .................... |
| D | 5902 hCoV-19/South Africa/Tygerberg_16/2020\|EPI_ISL_464 | .................... | .................... | .................... | .................... | .................... | .................... |
| D | 5903 hCoV-19/South Africa/Tygerberg_17/2020\|EPI_ISL_464 | .................... | .................... | .................... | .................... | .................... | .................... |
| D | 5904 hCoV-19/South Africa/Tygerberg_18/2020\|EPI_ISL_464 | .................... | .................... | .................... | .................... | .................... | .................... |
| D | 5905 hCoV-19/South Africa/Tygerberg_19/2020\|EPI_ISL_464 | .................... | .................... | .................... | .................... | .................... | .................... |
| D | 5906 hCoV-19/South Africa/Tygerberg_20/2020\|EPI_ISL_464 | .................... | .................... | .................... | .................... | .................... | .................... |
| D | 5907 hCoV-19/South Africa/Tygerberg_21/2020\|EPI_ISL_464 | .................... | .................... | .................... | .................... | .................... | .................... |
| D | 5908 hCoV-19/South Africa/Tygerberg_22/2020\|EPI_ISL_464 | .................... | .................... | .................... | .................... | .................... | .................... |
| D | 5909 hCoV-19/England/20180032304/2020\|EPI_ISL_465436\|20 | .................... | .................... | .................... | .................... | .................... | .................... |

| | | | | | | | | |
|---|---|---|---|---|---|---|---|---|
| D | 5910 hCoV-19/England/20180044804/2020\|EPI_ISL_465440\|20 | .................... | .................... | .................... | .................... | .................... | .................... | |
| D | 5911 hCoV-19/England/20182004604/2020\|EPI_ISL_465441\|20 | .................... | .................... | .................... | .................... | .................... | .................... | |
| D | 5912 hCoV-19/England/20182008204/2020\|EPI_ISL_465448\|20 | .................... | .................... | .................... | .................... | .................... | .................... | |
| D | 5913 hCoV-19/England/20182013104/2020\|EPI_ISL_465450\|20 | .................... | .................... | .................... | .................... | .................... | .................... | |
| D | 5914 hCoV-19/England/20184026604/2020\|EPI_ISL_465453\|20 | .................... | .................... | .................... | .................... | .................... | .................... | |
| D | 5915 hCoV-19/England/20184026804/2020\|EPI_ISL_465454\|20 | .................... | .................... | .................... | .................... | .................... | .................... | |
| D | 5916 hCoV-19/England/20184046404/2020\|EPI_ISL_465458\|20 | .................... | .................... | .................... | .................... | .................... | .................... | |
| D | 5917 hCoV-19/England/20186005504/2020\|EPI_ISL_465464\|20 | .................... | .................... | .................... | .................... | .................... | .................... | |
| D | 5918 hCoV-19/England/20186006704/2020\|EPI_ISL_465466\|20 | .................... | .................... | .................... | .................... | .................... | .................... | |
| D | 5919 hCoV-19/Canada/BC_04286257/2020\|EPI_ISL_466765\|202 | .................... | .................... | .................... | .................... | .................... | .................... | |
| D | 5920 hCoV-19/Canada/BC_04346268/2020\|EPI_ISL_466766\|202 | .................... | .A.................. | .................... | .................... | .................... | .................... | |
| D | 5921 hCoV-19/Canada/BC_04837165/2020\|EPI_ISL_466767\|202 | .................... | .................... | .................... | .................... | .................... | .................... | |
| D | 5922 hCoV-19/Canada/BC_04837932/2020\|EPI_ISL_466768\|202 | .................... | .................... | .................... | .................... | .................... | .................... | |
| D | 5923 hCoV-19/Canada/BC_04882357/2020\|EPI_ISL_466770\|202 | .................... | .................... | .................... | .................... | .................... | .................... | |
| D | 5924 hCoV-19/Canada/BC_04930828/2020\|EPI_ISL_466772\|202 | .................... | .................... | .................... | .................... | .................... | .................... | |
| D | 5925 hCoV-19/Canada/BC_04935664/2020\|EPI_ISL_466773\|202 | .................... | .................... | .................... | .................... | .................... | .................... | |
| D | 5926 hCoV-19/Canada/BC_04975041/2020\|EPI_ISL_466774\|202 | .................... | .................... | .................... | .................... | .................... | .................... | |
| D | 5927 hCoV-19/Canada/BC_05017368/2020\|EPI_ISL_466775\|202 | .................... | .................... | .................... | .................... | .................... | .................... | |
| D | 5928 hCoV-19/Canada/BC_05023412/2020\|EPI_ISL_466776\|202 | .................... | .................... | .................... | .................... | .................... | .................... | |
| D | 5929 hCoV-19/Canada/BC_05075366/2020\|EPI_ISL_466777\|202 | .................... | .................... | .................... | .................... | .................... | .................... | |
| D | 5930 hCoV-19/Canada/BC_05075882/2020\|EPI_ISL_466779\|202 | .................... | .................... | .................... | .................... | .................... | .................... | |
| D | 5931 hCoV-19/Canada/BC_05076038/2020\|EPI_ISL_466781\|202 | .................... | .................... | .................... | .................... | .................... | .................... | |
| D | 5932 hCoV-19/Canada/BC_05132390/2020\|EPI_ISL_466782\|202 | .................... | .................... | .................... | .................... | .................... | .................... | |
| D | 5933 hCoV-19/Canada/BC_05248019/2020\|EPI_ISL_466783\|202 | .................... | .................... | .................... | .................... | .................... | .................... | |
| D | 5934 hCoV-19/Canada/BC_05256838/2020\|EPI_ISL_466784\|202 | .................... | .................... | .................... | .................... | .................... | .................... | |
| D | 5935 hCoV-19/Canada/BC_05294838/2020\|EPI_ISL_466785\|202 | .................... | .................... | .................... | .................... | .................... | .................... | |
| D | 5936 hCoV-19/Canada/BC_05300317/2020\|EPI_ISL_466787\|202 | .................... | .................... | .................... | .................... | .................... | .................... | |
| D | 5937 hCoV-19/Canada/BC_05333597/2020\|EPI_ISL_466788\|202 | .................... | .................... | .................... | .................... | .................... | .................... | |
| D | 5938 hCoV-19/Canada/BC_05740462/2020\|EPI_ISL_466789\|202 | .................... | .................... | .................... | .................... | .................... | .................... | |
| D | 5939 hCoV-19/Canada/BC_05941640/2020\|EPI_ISL_466792\|202 | .................... | .................... | .................... | .................... | .................... | .................... | |
| D | 5940 hCoV-19/Canada/BC_05941876/2020\|EPI_ISL_466793\|202 | .................... | .................... | .................... | .................... | .................... | .................... | |
| D | 5941 hCoV-19/Canada/BC_05941974/2020\|EPI_ISL_466794\|202 | .................... | .................... | .................... | .................... | .................... | .................... | |
| D | 5942 hCoV-19/Canada/BC_05942483/2020\|EPI_ISL_466795\|202 | .................... | .................... | .................... | .................... | .................... | .................... | |
| D | 5943 hCoV-19/Canada/BC_05943217/2020\|EPI_ISL_466796\|202 | .................... | .................... | .................... | .................... | .................... | .................... | |
| D | 5944 hCoV-19/USA/CT-Yale-278/2020\|EPI_ISL_469201\|2020-0 | .................... | .................... | .................... | .................... | .................... | .................... | |
| D | 5945 hCoV-19/USA/CT-Yale-280/2020\|EPI_ISL_469203\|2020-0 | .................... | .................... | .................... | .................... | .................... | .................... | |
| D | 5946 hCoV-19/USA/CT-Yale-282/2020\|EPI_ISL_469204\|2020-0 | .................... | ...................A | .................... | .................... | .................... | .................... | 1 |
| D | 5947 hCoV-19/USA/CT-Yale-286/2020\|EPI_ISL_469208\|2020-0 | .................... | .................... | .................... | .................... | .................... | .................... | |
| D | 5948 hCoV-19/Canada/BC_18173416/2020\|EPI_ISL_469209\|202 | .................... | .................... | .................... | .................... | .................... | .................... | |
| D | 5949 hCoV-19/Canada/BC_18604452/2020\|EPI_ISL_469210\|202 | .................... | .................... | .................... | .................... | .................... | .................... | |
| D | 5950 hCoV-19/Canada/BC_18717764/2020\|EPI_ISL_469211\|202 | .................... | .................... | .................... | .................... | .................... | .................... | |
| D | 5951 hCoV-19/Canada/BC_18839283/2020\|EPI_ISL_469212\|202 | .................... | .................... | .................... | .................... | .................... | .................... | |
| D | 5952 hCoV-19/Canada/BC_19071912/2020\|EPI_ISL_469213\|202 | .................... | .................... | .................... | .................... | .................... | .................... | |
| D | 5953 hCoV-19/Canada/BC_19072825/2020\|EPI_ISL_469214\|202 | .................... | .................... | .................... | .................... | .................... | .................... | |
| D | 5954 hCoV-19/Canada/BC_19072819/2020\|EPI_ISL_469215\|202 | .................... | .................... | .................... | .................... | .................... | .................... | |
| D | 5955 hCoV-19/Canada/BC_19100589/2020\|EPI_ISL_469216\|202 | .................... | .................... | .................... | .................... | .................... | .................... | |
| D | 5956 hCoV-19/Canada/BC_19133870/2020\|EPI_ISL_469218\|202 | .................... | .................... | .................... | .................... | .................... | .................... | |
| D | 5957 hCoV-19/Canada/BC_19677621/2020\|EPI_ISL_469222\|202 | .................... | .................... | .................... | .................... | .................... | .................... | |
| D | 5958 hCoV-19/Canada/BC_19803546/2020\|EPI_ISL_469223\|202 | .................... | .................... | .................... | .................... | .................... | .................... | |
| D | 5959 hCoV-19/Canada/NL_82/2020\|EPI_ISL_469224\|2020-03-2 | .................... | .................... | .................... | .................... | .................... | .................... | |
| D | 5960 hCoV-19/Canada/NL_83/2020\|EPI_ISL_469225\|2020-03-2 | .................... | .................... | .................... | .................... | .................... | .................... | |
| D | 5961 hCoV-19/Canada/NL_84/2020\|EPI_ISL_469226\|2020-03-2 | .................... | .................... | .................... | .................... | .................... | .................... | |
| D | 5962 hCoV-19/Canada/NL_85/2020\|EPI_ISL_469227\|2020-03-2 | .................... | .................... | .................... | .................... | .................... | .................... | |
| D | 5963 hCoV-19/Canada/NL_86/2020\|EPI_ISL_469228\|2020-03-2 | .................... | .................... | .................... | .................... | .................... | .................... | |
| D | 5964 hCoV-19/Singapore/72/2020\|EPI_ISL_469274\|2020-04-0 | .................... | .................... | .................... | .................... | .................... | .................... | |
| D | 5965 hCoV-19/Egypt/CUNCI-HGC002-2/2020\|EPI_ISL_469275\|2 | .................... | .....R.............. | .................... | .................... | .................... | .................... | |
| D | 5966 hCoV-19/United Arab Emirates/L0287/2020\|EPI_ISL_46 | .................... | .................... | .................... | .................... | .................... | .................... | |
| D | 5967 hCoV-19/United Arab Emirates/L1189/2020\|EPI_ISL_46 | .................... | .................... | .................... | .................... | .................... | .................... | |
| D | 5968 hCoV-19/United Arab Emirates/L4711/2020\|EPI_ISL_46 | .................... | .................... | .................... | .................... | .................... | .................... | |
| D | 5969 hCoV-19/United Arab Emirates/L5857/2020\|EPI_ISL_46 | .................... | .................... | .................... | .................... | .................... | .................... | |
| D | 5970 hCoV-19/United Arab Emirates/L6841/2020\|EPI_ISL_46 | .................... | .................... | .................... | .................... | .................... | .................... | |
| D | 5971 hCoV-19/United Arab Emirates/L9119/2020\|EPI_ISL_46 | .................... | .................... | .................... | .................... | .................... | .................... | |
| D | 5972 hCoV-19/France/IDF-211-80/2020\|EPI_ISL_469282\|2020 | .................... | .................... | .................... | .................... | .................... | .................... | |
| D | 5973 hCoV-19/France/IDF-221-61/2020\|EPI_ISL_469283\|2020 | .................... | .................... | .................... | .................... | .................... | .................... | |
| D | 5974 hCoV-19/France/IDF-220-95/2020\|EPI_ISL_469284\|2020 | .................... | .................... | .................... | .................... | .................... | .................... | |
| D | 5975 hCoV-19/Bangladesh/BCSIR-NILMRC-094/2020\|EPI_ISL_4 | .................... | .................... | .................... | .................... | .................... | .................... | |
| D | 5976 hCoV-19/Japan/Donner29/2020\|EPI_ISL_469287\|2020-05 | .................... | .................... | .................... | .................... | .................... | .................... | |
| D | 5977 hCoV-19/Japan/Donner30/2020\|EPI_ISL_469288\|2020-05 | .................... | .................... | .................... | .................... | .................... | .................... | |
| D | 5978 hCoV-19/Japan/Donner31/2020\|EPI_ISL_469289\|2020-04 | .................... | .................... | .................... | .................... | .................... | .................... | |
| D | 5979 hCoV-19/Japan/Donner36/2020\|EPI_ISL_469294\|2020-05 | .................... | .................... | .................... | .................... | .................... | .................... | |
| D | 5980 hCoV-19/Japan/Donner37/2020\|EPI_ISL_469295\|2020-05 | .................... | .................... | .................... | .................... | .................... | .................... | |
| D | 5981 hCoV-19/Bangladesh/BCSIR-NILMRC-095/2020\|EPI_ISL_4 | .................... | .................... | .................... | .................... | .................... | .................... | |
| D | 5982 hCoV-19/England/CAMB-1AA69C/2020\|EPI_ISL_470200\|20 | .................... | .................... | .................... | .................... | .................... | .................... | |
| D | 5983 hCoV-19/England/CAMB-1AB25B/2020\|EPI_ISL_470201\|20 | .................... | .................... | .................... | .................... | .................... | .................... | |
| D | 5984 hCoV-19/England/CAMB-1AAB9D/2020\|EPI_ISL_470202\|20 | .................... | .................... | .................... | .................... | .................... | .................... | |
| D | 5985 hCoV-19/England/CAMB-1AB8D1/2020\|EPI_ISL_470203\|20 | .................... | .................... | .................... | .................... | .................... | .................... | |
| D | 5986 hCoV-19/England/CAMB-1AB3C1/2020\|EPI_ISL_470204\|20 | .................... | .................... | .................... | .................... | .................... | .................... | |
| D | 5987 hCoV-19/England/CAMB-1ABE0C/2020\|EPI_ISL_470205\|20 | .................... | .................... | .................... | .................... | .................... | .................... | |

| | | | | | | | | | |
|---|---|---|---|---|---|---|---|---|---|
| D | 5988 | hCoV-19/England/CAMB-1AA650/2020\|EPI_ISL_470206\|20 | ···················· | ···················· | ···················· | ···················· | ···················· | ···················· | |
| D | 5989 | hCoV-19/England/CAMB-1AB552/2020\|EPI_ISL_470208\|20 | ···················· | ···················· | ···················· | ···················· | ···················· | ···················· | |
| D | 5990 | hCoV-19/England/CAMB-1AA7C6/2020\|EPI_ISL_470209\|20 | ···················· | ···················· | ···················· | ···················· | ···················· | ···················· | |
| D | 5991 | hCoV-19/England/CAMB-1AA78A/2020\|EPI_ISL_470229\|20 | ···················· | ···················· | ···················· | ···················· | ······T············· | ···················· | |
| D | 5992 | hCoV-19/England/CAMB-1AB859/2020\|EPI_ISL_470231\|20 | ···················· | ···················· | ···················· | ···················· | ···················· | ···················· | |
| D | 5993 | hCoV-19/England/CAMB-1AB2C4/2020\|EPI_ISL_470232\|20 | ···················· | ·A·················· | ···················· | ···················· | ···················· | ···················· | |
| D | 5994 | hCoV-19/England/CAMB-1AA73F/2020\|EPI_ISL_470233\|20 | ···················· | ···················· | ···················· | ···················· | ···················· | ···················· | |
| D | 5995 | hCoV-19/England/CAMB-1AAA90/2020\|EPI_ISL_470235\|20 | ···················· | ···················· | ···················· | ···················· | ···················· | ···················· | |
| D | 5996 | hCoV-19/England/CAMB-1AB0F7/2020\|EPI_ISL_470236\|20 | ···················· | ···················· | ···················· | ···················· | ···················· | ···················· | |
| D | 5997 | hCoV-19/England/CAMB-1AA6C9/2020\|EPI_ISL_470237\|20 | ···················· | ···················· | ···················· | ···················· | ···················· | ···················· | |
| D | 5998 | hCoV-19/England/CAMB-1AAB60/2020\|EPI_ISL_470239\|20 | ···················· | ···················· | ···················· | ···················· | ···················· | ···················· | |
| D | 5999 | hCoV-19/England/CAMB-1AB21F/2020\|EPI_ISL_470241\|20 | ···················· | ···················· | ···················· | ···················· | ···················· | ···················· | |
| D | 6000 | hCoV-19/England/CAMB-1AA9B1/2020\|EPI_ISL_470244\|20 | ···················· | ···················· | ···················· | ···················· | ···················· | ···················· | |
| D | 6001 | hCoV-19/England/CAMB-1AB4DD/2020\|EPI_ISL_470245\|20 | ···················· | ···················· | ···················· | ···················· | ···················· | ···················· | |
| D | 6002 | hCoV-19/England/CAMB-1ABCB7/2020\|EPI_ISL_470246\|20 | ···················· | ···················· | ···················· | ···················· | ···················· | ···················· | |
| D | 6003 | hCoV-19/England/CAMB-1ABCF3/2020\|EPI_ISL_470248\|20 | ···················· | ···················· | ···················· | ···················· | ···················· | ···················· | |
| D | 6004 | hCoV-19/England/CAMB-1ABD2D/2020\|EPI_ISL_470249\|20 | ···················· | ···················· | ···················· | ···················· | ···················· | ···················· | |
| D | 6005 | hCoV-19/England/CAMB-1AACF4/2020\|EPI_ISL_470250\|20 | ···················· | ···················· | ···················· | ···················· | ···················· | ···················· | |
| D | 6006 | hCoV-19/England/CAMB-1AA957/2020\|EPI_ISL_470251\|20 | ···················· | ···················· | ···················· | ···················· | ···················· | ···················· | |
| D | 6007 | hCoV-19/England/20236092104/2020\|EPI_ISL_471510\|20 | ···················· | ···················· | ···················· | ···················· | ···················· | ···················· | |
| D | 6008 | hCoV-19/England/20238001204/2020\|EPI_ISL_471512\|20 | ···················· | ···················· | ···················· | ···················· | ···················· | ···················· | |
| D | 6010 | hCoV-19/England/20238073804/2020\|EPI_ISL_471516\|20 | ···················· | ···················· | ···················· | ···················· | ···················· | ···················· | |
| D | 6011 | hCoV-19/England/20239016504/2020\|EPI_ISL_471517\|20 | ···················· | ···················· | ···················· | ···················· | ···················· | ···················· | |
| D | 6014 | hCoV-19/England/20242090304/2020\|EPI_ISL_471521\|20 | ···················· | ···················· | ···················· | ···················· | ···················· | ···················· | |
| D | 6015 | hCoV-19/England/20244086004/2020\|EPI_ISL_471524\|20 | ···················· | ···················· | ···················· | ···················· | ···················· | ···················· | |
| D | 6017 | hCoV-19/Czech Republic/NRL_1751/2020\|EPI_ISL_47152 | ···················· | ···················· | ···················· | ···················· | ···················· | ···················· | |
| D | 6018 | hCoV-19/Czech Republic/NRL_1885/2020\|EPI_ISL_47153 | ···················· | ···················· | ···················· | ···················· | ···················· | ···················· | |
| D | 6019 | hCoV-19/Brazil/SP-47/2020\|EPI_ISL_471539\|2020-03-1 | ···················· | ···················· | ···················· | ···················· | ···················· | ···················· | |
| D | 6020 | hCoV-19/Brazil/SP-523/2020\|EPI_ISL_471562\|2020-04- | ···················· | ···················· | ···················· | ···················· | ···················· | ···················· | |
| D | 6021 | hCoV-19/Brazil/SP-524/2020\|EPI_ISL_471581\|2020-04- | ···················· | ···················· | ···················· | ···················· | ···················· | ···················· | |
| D | 6022 | hCoV-19/Brazil/SP-525/2020\|EPI_ISL_471582\|2020-04- | ········T··········· | ···················· | ···················· | ···················· | ···················· | ···················· | |
| D | 6023 | hCoV-19/Wales/PHWC-15E1E1/2020\|EPI_ISL_472800\|2020 | ···················· | ···················· | ···················· | ···················· | ···················· | ···················· | |
| D | 6024 | hCoV-19/Wales/PHWC-15E275/2020\|EPI_ISL_472801\|2020 | ···················· | ···················· | ···················· | ···················· | ···················· | ···················· | |
| D | 6025 | hCoV-19/Wales/PHWC-15E327/2020\|EPI_ISL_472805\|2020 | ···················· | ···················· | ···················· | ···················· | ···················· | ···················· | |
| D | 6026 | hCoV-19/Wales/PHWC-15E372/2020\|EPI_ISL_472808\|2020 | ···················· | ···················· | ···················· | ···················· | ···················· | ···················· | |
| D | 6027 | hCoV-19/Wales/PHWC-15E381/2020\|EPI_ISL_472809\|2020 | ···················· | ···················· | ···················· | ···················· | ···················· | ···················· | |
| D | 6028 | hCoV-19/Wales/PHWC-15E3AF/2020\|EPI_ISL_472810\|2020 | ···················· | ···················· | ···················· | ···················· | ···················· | ···················· | |
| D | 6029 | hCoV-19/Wales/PHWC-15E3DC/2020\|EPI_ISL_472812\|2020 | ···················· | ···················· | ···················· | ···················· | ···················· | ···················· | |
| D | 6030 | hCoV-19/Wales/PHWC-15E3EB/2020\|EPI_ISL_472813\|2020 | ···················· | ···················· | ···················· | ···················· | ···················· | ···················· | |
| D | 6031 | hCoV-19/Wales/PHWC-15E406/2020\|EPI_ISL_472814\|2020 | ···················· | ···················A | ···················· | ···················· | ···················· | ···················· | 1 |
| D | 6032 | hCoV-19/Wales/PHWC-15E415/2020\|EPI_ISL_472815\|2020 | ···················· | ···················· | ···················· | ···················· | ···················· | ···················· | |
| D | 6033 | hCoV-19/Wales/PHWC-15E4F7/2020\|EPI_ISL_472818\|2020 | ···················· | ···················· | ···················· | ···················· | ···················· | ···················· | |
| D | 6034 | hCoV-19/Wales/PHWC-15E58B/2020\|EPI_ISL_472820\|2020 | ···················· | ····A··············· | ···················· | ···················· | ···················· | ···················· | |
| D | 6035 | hCoV-19/Wales/PHWC-15E59A/2020\|EPI_ISL_472821\|2020 | ···················· | ···················· | ···················· | ···················· | ···················· | ···················· | |
| D | 6036 | hCoV-19/Wales/PHWC-15E61F/2020\|EPI_ISL_472823\|2020 | ···················· | ···················· | ···················· | ···················· | ···················· | ···················· | |
| D | 6037 | hCoV-19/Wales/PHWC-15E66A/2020\|EPI_ISL_472824\|2020 | ···················· | ···················· | ···················· | ···················· | ···················· | ···················· | |
| D | 6038 | hCoV-19/Wales/PHWC-15E697/2020\|EPI_ISL_472825\|2020 | ···················· | ···················· | ···················· | ···················· | ···················· | ···················· | |
| D | 6039 | hCoV-19/Wales/PHWC-15E794/2020\|EPI_ISL_472830\|2020 | ···················· | ···················· | ···················· | ···················· | ····T··············· | ···················· | |
| D | 6040 | hCoV-19/Wales/PHWC-15E7D0/2020\|EPI_ISL_472831\|2020 | ···················· | ···················· | ···················· | ···················· | ···················· | ···················· | |
| D | 6041 | hCoV-19/Wales/PHWC-15E7FE/2020\|EPI_ISL_472833\|2020 | ···················· | ···················· | ···················· | ···················· | ···················· | ···················· | |
| D | 6042 | hCoV-19/Wales/PHWC-15E80A/2020\|EPI_ISL_472834\|2020 | ···················· | ···················· | ···················· | ···················· | ···················· | ···················· | |
| D | 6043 | hCoV-19/Wales/PHWC-15E819/2020\|EPI_ISL_472835\|2020 | ···················· | ···················· | ···················· | ···················· | ···················· | ···················· | |
| D | 6044 | hCoV-19/Wales/PHWC-15E855/2020\|EPI_ISL_472838\|2020 | ···················· | ···················· | ···················· | ···················· | ···················· | ···················· | |
| D | 6045 | hCoV-19/Wales/PHWC-15E873/2020\|EPI_ISL_472839\|2020 | ···················· | ···················· | ···················· | ···················· | ···················· | ···················· | |
| D | 6046 | hCoV-19/Wales/PHWC-15E970/2020\|EPI_ISL_472843\|2020 | ···················· | ····A··············· | ···················· | ···················· | ···················· | ···················· | |
| D | 6047 | hCoV-19/Wales/PHWC-15FCB2/2020\|EPI_ISL_472844\|2020 | ···················· | ···················· | ···················· | ···················· | ···················· | ···················· | |
| D | 6048 | hCoV-19/Wales/PHWC-15FCD0/2020\|EPI_ISL_472845\|2020 | ···················· | ···················· | ···················· | ···················· | ···················· | ···················· | |
| D | 6049 | hCoV-19/Wales/PHWC-15FCEF/2020\|EPI_ISL_472846\|2020 | ···················· | ···················· | ···················· | ···················· | ···················· | ···················· | |
| D | 6050 | hCoV-19/Wales/PHWC-15FD28/2020\|EPI_ISL_472848\|2020 | ···················· | ···················· | ···················· | ···················· | ···················· | ···················· | |
| D | 6051 | hCoV-19/Wales/PHWC-15FD55/2020\|EPI_ISL_472850\|2020 | ···················· | ···················· | ···················· | ···················· | ···················· | ···················· | |
| D | 6052 | hCoV-19/Wales/PHWC-15FD64/2020\|EPI_ISL_472851\|2020 | ···················· | ···················· | ···················· | ···················· | ···················· | ···················· | |
| D | 6053 | hCoV-19/Wales/PHWC-15FD91/2020\|EPI_ISL_472854\|2020 | ···················· | ···················· | ···················· | ···················· | ···················· | ···················· | |
| D | 6054 | hCoV-19/Wales/PHWC-15FDDD/2020\|EPI_ISL_472856\|2020 | ···················· | ···················· | ···················· | ···················· | ···················· | ···················· | |
| D | 6055 | hCoV-19/Wales/PHWC-15FDEC/2020\|EPI_ISL_472857\|2020 | ···················· | ···················· | ···················· | ···················· | ···················· | ···················· | |
| D | 6056 | hCoV-19/Wales/PHWC-15FDFB/2020\|EPI_ISL_472858\|2020 | ···················· | ···················· | ···················· | ···················· | ···················· | ···················· | |
| D | 6057 | hCoV-19/Wales/PHWC-15FE25/2020\|EPI_ISL_472861\|2020 | ···················· | ···················· | ···················· | ···················· | ···················· | ···················· | |
| D | 6058 | hCoV-19/Wales/PHWC-160085/2020\|EPI_ISL_472890\|2020 | ···················· | ···················· | ···················· | ···················· | ···················· | ···················· | |
| D | 6059 | hCoV-19/Wales/PHWC-1600A3/2020\|EPI_ISL_472892\|2020 | ···················· | ···················· | ···················· | ···················· | ···················· | ···················· | |
| D | 6060 | hCoV-19/Wales/PHWC-1600D0/2020\|EPI_ISL_472895\|2020 | ···················· | ···················· | ···················· | ···················· | ···················· | ···················· | |
| D | 6061 | hCoV-19/Wales/PHWC-1600FE/2020\|EPI_ISL_472896\|2020 | ···················· | ···················· | ···················· | ···················· | ···················· | ···················· | |
| D | 6062 | hCoV-19/Wales/PHWC-160119/2020\|EPI_ISL_472898\|2020 | ···················· | ···················· | ···················· | ···················· | ···················· | ···················· | |
| D | 6063 | hCoV-19/Wales/PHWC-160146/2020\|EPI_ISL_472899\|2020 | ···················· | ···················· | ···················· | ···················· | ···················· | ···················· | |
| D | 6064 | hCoV-19/Belgium/UGent-110/2020\|EPI_ISL_475070\|2020 | ···················· | ···················· | ···················· | ···················· | ···················· | ···················· | |
| D | 6066 | hCoV-19/Belgium/UGent-112/2020\|EPI_ISL_475072\|2020 | ···················· | ···················· | ···················· | ···················· | ···················· | ···················· | |
| D | 6067 | hCoV-19/Belgium/UGent-113/2020\|EPI_ISL_475073\|2020 | ···················· | ···················· | ···················· | ···················· | ···················· | ···················· | |
| D | 6068 | hCoV-19/Belgium/UGent-114/2020\|EPI_ISL_475074\|2020 | ···················· | ···················· | ···················· | ···················· | ···················· | ···················· | |
| D | 6070 | hCoV-19/Belgium/UGent-116/2020\|EPI_ISL_475076\|2020 | ···················· | ···················· | ···················· | ···················· | ···················· | ···················· | |
| D | 6071 | hCoV-19/Belgium/UGent-117/2020\|EPI_ISL_475077\|2020 | ···················· | ···················· | ···················· | ···················· | ···················· | ···················· | |

| | | | | | | | | |
|---|---|---|---|---|---|---|---|---|
| D | 6076 hCoV-19/Belgium/UGent-122/2020\|EPI_ISL_475082\|2020 | ···················· | ···················· | ···················· | ···················· | ···················· | ···················· | |
| D | 6077 hCoV-19/Bangladesh/BCSIR-NILMRC-101/2020\|EPI_ISL_4 | ···················· | ···················· | ···················· | ···················· | ···················· | ···················· | |
| D | 6078 hCoV-19/Bangladesh/BCSIR-NILMRC-103/2020\|EPI_ISL_4 | ···················· | ···················· | ···················· | ···················· | ···················· | ···················· | |
| D | 6079 hCoV-19/Sweden/20-08174/2020\|EPI_ISL_475085\|2020-0 | ···················· | ·A·················· | ···················· | ···················· | ···················· | ···················· | |
| D | 6080 hCoV-19/Sweden/20-50327/2020\|EPI_ISL_475086\|2020-0 | ···················· | ···················· | ···················· | ···················· | ···················· | ···················· | |
| D | 6081 hCoV-19/Sweden/20-50335/2020\|EPI_ISL_475087\|2020-0 | ···················· | ···················· | ···················· | ···················· | ···················· | ···················· | |
| D | 6082 hCoV-19/Sweden/20-50336/2020\|EPI_ISL_475088\|2020-0 | ···················· | ···················· | ···················· | ···················· | ···················· | ···················· | |
| D | 6083 hCoV-19/Sweden/20-50337/2020\|EPI_ISL_475089\|2020-0 | ···················· | ···················· | ···················· | ···················· | ···················· | ···················· | |
| D | 6084 hCoV-19/Sweden/20-50341/2020\|EPI_ISL_475090\|2020-0 | ···················· | ···················· | ···················· | ···················· | ···················· | ···················· | |
| D | 6085 hCoV-19/Sweden/20-50342/2020\|EPI_ISL_475091\|2020-0 | ···················· | ···················· | ···················· | ···················· | ···················· | ···················· | |
| D | 6086 hCoV-19/Sweden/20-08209/2020\|EPI_ISL_475092\|2020-0 | ···················· | ···················· | ···················· | ···················· | ···················· | ···················· | |
| D | 6087 hCoV-19/Sweden/20-08210/2020\|EPI_ISL_475093\|2020-0 | ···················· | ···················· | ···················· | ···················· | ···················· | ···················· | |
| D | 6088 hCoV-19/Sweden/20-08211/2020\|EPI_ISL_475094\|2020-0 | ···················· | ···················· | ···················· | ···················· | ···················· | ···················· | |
| D | 6089 hCoV-19/Sweden/20-50362/2020\|EPI_ISL_475095\|2020-0 | ···················· | ···················· | ···················· | ···················· | ···················· | ···················· | |
| D | 6090 hCoV-19/Sweden/20-08317/2020\|EPI_ISL_475096\|2020-0 | ···················· | ···················· | ···················· | ···················· | ···················· | ···················· | |
| D | 6091 hCoV-19/Sweden/20-08311/2020\|EPI_ISL_475097\|2020-0 | ···················· | ···················· | ···················· | ···················· | ···················· | ···················· | |
| D | 6092 hCoV-19/Sweden/20-08328/2020\|EPI_ISL_475098\|2020-0 | ···················· | ···················· | ···················· | ···················· | ···················· | ···················· | |
| D | 6093 hCoV-19/Sweden/20-08387/2020\|EPI_ISL_475099\|2020-0 | ···················· | ···················· | ···················· | ···················· | ···················· | ···················· | |
| D | 6095 hCoV-19/Switzerland/BE-ETHZ-140068/2020\|EPI_ISL_47 | ···················· | ···················· | ···················· | ···················· | ···················· | ···················· | |
| D | 6096 hCoV-19/Switzerland/ZH-ETHZ-140069/2020\|EPI_ISL_47 | ···················· | ···················· | ···················· | ···················· | ······M············· | W··················· | |
| D | 6097 hCoV-19/Switzerland/ZH-ETHZ-140070/2020\|EPI_ISL_47 | ···················· | ···················· | ···················· | ···················· | ······S············· | ···················· | |
| D | 6098 hCoV-19/Switzerland/ZH-ETHZ-140071/2020\|EPI_ISL_47 | ··················Y· | ···················· | ···················· | ···················· | ······S············· | ···················· | 1 |
| D | 6099 hCoV-19/Switzerland/UR-ETHZ-140073/2020\|EPI_ISL_47 | ···················· | ···················· | ···················· | ···················· | ···················· | ···················· | |
| D | 6100 hCoV-19/Switzerland/UR-ETHZ-140074/2020\|EPI_ISL_47 | ···················· | ···················· | ···················· | ···················· | ······S···M········· | ···················· | |
| D | 6101 hCoV-19/Switzerland/SO-ETHZ-140075/2020\|EPI_ISL_47 | ···················· | ···················· | ···················· | ···················· | ······V············· | ···················· | |
| D | 6102 hCoV-19/Switzerland/GE-ETHZ-140077/2020\|EPI_ISL_47 | ···················· | ···················· | ···················· | ···················· | ···················· | ···················· | |
| D | 6103 hCoV-19/Switzerland/SO-ETHZ-140079/2020\|EPI_ISL_47 | ···················· | ···················· | ···················· | ···················· | ···················· | ···················· | |
| D | 6104 hCoV-19/Switzerland/ZH-ETHZ-140083/2020\|EPI_ISL_47 | ···················· | ···················· | ···················· | ···················· | ···················· | ···················· | |
| D | 6105 hCoV-19/Switzerland/BE-ETHZ-140084/2020\|EPI_ISL_47 | ···················· | ···················· | ···················· | ···················· | ···················· | ···················· | |
| D | 6106 hCoV-19/Switzerland/BE-ETHZ-140085/2020\|EPI_ISL_47 | ···················· | ···················· | ···················· | ···················· | ···················· | ···················· | |
| D | 6108 hCoV-19/Switzerland/GE-ETHZ-140087/2020\|EPI_ISL_47 | ···················· | ···················A | ···················· | ···················· | ···················· | ···················· | 1 |
| D | 6109 hCoV-19/Switzerland/BL-ETHZ-140089/2020\|EPI_ISL_47 | ···················· | ···················· | ···················· | ···················· | ···················· | ···················· | |
| D | 6110 hCoV-19/Switzerland/GR-ETHZ-140090/2020\|EPI_ISL_47 | ···················· | ···················· | ···················· | ···················· | ······M············· | ···················· | |
| D | 6111 hCoV-19/Switzerland/ZH-ETHZ-140091/2020\|EPI_ISL_47 | ···················· | ···················· | ···················· | ···················· | ···················· | ···················· | |
| D | 6112 hCoV-19/Switzerland/GE-ETHZ-140092/2020\|EPI_ISL_47 | ···················· | ···················· | ···················· | ···················· | ······S············· | ···················· | |
| D | 6113 hCoV-19/Sweden/20-51578/2020\|EPI_ISL_476135\|2020-0 | ···················· | ···················· | ···················· | ···················· | ···················· | ···················· | |
| D | 6114 hCoV-19/Sweden/20-51636/2020\|EPI_ISL_476136\|2020-0 | ···················· | ···················· | ···················· | ···················· | ···················· | ···················· | |
| D | 6115 hCoV-19/Sweden/20-51681/2020\|EPI_ISL_476137\|2020-0 | ···················· | ···················· | ···················· | ···················· | ···················· | ···················· | |
| D | 6116 hCoV-19/Sweden/20-51759/2020\|EPI_ISL_476138\|2020-0 | ···················· | ···················· | ···················· | ···················· | ···················· | ···················· | |
| D | 6117 hCoV-19/Sweden/20-02114/2020\|EPI_ISL_476139\|2020-0 | ···················· | ···················· | ···················· | ···················· | ···················· | ···················· | |
| D | 6118 hCoV-19/Sweden/20-08362/2020\|EPI_ISL_476142\|2020-0 | ···················· | ···················· | ···················· | ···················· | ···················· | ···················· | |
| D | 6119 hCoV-19/Sweden/20-08608/2020\|EPI_ISL_476144\|2020-0 | ···················· | ···················· | ···················· | ···················· | ···················· | ···················· | |
| D | 6120 hCoV-19/Sweden/20-08865/2020\|EPI_ISL_476145\|2020-0 | ···················· | ···················· | ···················· | ···················· | ···················· | ···················· | |
| D | 6121 hCoV-19/Sweden/20-08866/2020\|EPI_ISL_476146\|2020-0 | ···················· | ···················· | ···················· | ···················· | ···················· | ···················· | |
| D | 6122 hCoV-19/Sweden/20-08869/2020\|EPI_ISL_476147\|2020-0 | ···················· | ···················· | ···················· | ···················· | ···················· | ···················· | |
| D | 6123 hCoV-19/England/OXON-AF856/2020\|EPI_ISL_478777\|202 | ···················· | ···················· | ···················· | ···················· | ···················· | ···················· | |
| D | 6124 hCoV-19/England/OXON-AF865/2020\|EPI_ISL_478778\|202 | ···················· | ···················· | ···················· | ···················· | ···················· | ···················· | |
| D | 6125 hCoV-19/England/OXON-AF874/2020\|EPI_ISL_478779\|202 | ···················· | ···················· | ···················· | ···················· | ···················· | ···················· | |
| D | 6126 hCoV-19/England/OXON-AF883/2020\|EPI_ISL_478780\|202 | ···················· | ···················· | ···················· | ···················· | ···················· | ···················· | |
| D | 6127 hCoV-19/England/OXON-AF8A1/2020\|EPI_ISL_478781\|202 | ···················· | ···················· | ···················· | ···················· | ···················· | ···················· | |
| D | 6128 hCoV-19/England/OXON-AF8CF/2020\|EPI_ISL_478782\|202 | ···················· | ···················· | ···················· | ···················· | ···················· | ···················· | |
| D | 6129 hCoV-19/England/OXON-AF8DE/2020\|EPI_ISL_478783\|202 | ···················· | ···················· | ···················· | ···················· | ···················· | ···················· | |
| D | 6130 hCoV-19/England/OXON-AF908/2020\|EPI_ISL_478784\|202 | ···················· | ···················· | ···················· | ···················· | ···················· | ···················· | |
| D | 6131 hCoV-19/England/OXON-AF935/2020\|EPI_ISL_478786\|202 | ···················· | ···················· | ···················· | ···················· | ···················· | ···················· | |
| D | 6132 hCoV-19/England/OXON-AF953/2020\|EPI_ISL_478787\|202 | ···················· | ···················· | ···················· | ···················· | ···················· | ···················· | |
| D | 6133 hCoV-19/England/OXON-AF9BD/2020\|EPI_ISL_478790\|202 | ···T················ | ···················· | ···················· | ···················· | ···················· | ···················· | |
| D | 6134 hCoV-19/England/OXON-AF9CC/2020\|EPI_ISL_478791\|202 | ···················· | ···················· | ···················· | ···················· | ···················· | ···················· | |
| D | 6135 hCoV-19/England/OXON-AF9DB/2020\|EPI_ISL_478792\|202 | ···················· | ···················· | ···················· | ···················· | ···················· | ···················· | |
| D | 6136 hCoV-19/England/OXON-AF9EA/2020\|EPI_ISL_478793\|202 | ···················· | ···················· | ···················· | ···················· | ···················· | ···················· | |
| D | 6137 hCoV-19/England/OXON-AFA05/2020\|EPI_ISL_478794\|202 | ···················· | ···················· | ···················· | ···················· | ···················· | ···················· | |
| D | 6138 hCoV-19/England/OXON-AFA14/2020\|EPI_ISL_478795\|202 | ···················· | ···················· | ···················· | ···················· | ···················· | ···················· | |
| D | 6139 hCoV-19/England/OXON-AFA23/2020\|EPI_ISL_478796\|202 | ···················· | ···················· | ···················· | ···················· | ···················· | ···················· | |
| D | 6140 hCoV-19/England/OXON-AFA32/2020\|EPI_ISL_478797\|202 | ···················· | ···················· | ···················· | ···················· | ···················· | ···················· | |
| D | 6141 hCoV-19/England/OXON-AFA41/2020\|EPI_ISL_478798\|202 | ···················· | ···················· | ···················· | ···················· | ···················· | ···················· | |
| D | 6142 hCoV-19/England/OXON-AFA50/2020\|EPI_ISL_478799\|202 | ···················· | ···················· | ···················· | ···················· | ···················· | ···················· | |
| D | 6143 hCoV-19/cat/Hong Kong/20-04236/2020\|EPI_ISL_759858 | ···················· | ···················· | ···················· | ···················· | ···················· | ···················· | |
| D | 6144 hCoV-19/Hong Kong/VM20005127/2020\|EPI_ISL_759859\|2 | ···················· | ···················· | ···················· | ···················· | ···················· | ···················· | |
| D | 6145 hCoV-19/South Africa/Tygerberg_26/2020\|EPI_ISL_464 | ···················· | ···················· | ···················· | ···················· | ···················· | ···················· | |
| D | 6146 hCoV-19/South Africa/Tygerberg_27/2020\|EPI_ISL_464 | ···················· | ···················· | ···················· | ···················· | ···················· | ···················· | |
| D | 6147 hCoV-19/South Africa/Tygerberg_34/2020\|EPI_ISL_464 | ···················· | ···················· | ···················· | ···················· | ···················· | ···················· | |
| D | 6148 hCoV-19/South Africa/Tygerberg_44/2020\|EPI_ISL_464 | ···················· | ···················· | ···················· | ···················· | ···················· | ···················· | |
| D | 6149 hCoV-19/South Africa/Tygerberg_46/2020\|EPI_ISL_464 | ···················· | ···················· | ···················· | ···················· | ···················· | ···················· | |
| D | 6150 hCoV-19/South Africa/Tygerberg_48/2020\|EPI_ISL_464 | ···················· | ···················· | ···················· | ···················· | ···················· | ···················· | |
| D | 6151 hCoV-19/Bangladesh/BCSIR-NILMRC-087/2020\|EPI_ISL_4 | ···················· | ···················· | ···················· | ···················· | ···················· | ···················· | |
| D | 6152 hCoV-19/Bangladesh/BCSIR-NILMRC-084/2020\|EPI_ISL_4 | ···················· | ···················· | ···················· | ···················· | ···················· | ···················· | |
| D | 6153 hCoV-19/Bangladesh/BCSIR-NILMRC-083/2020\|EPI_ISL_4 | ···················· | ···················· | ···················· | ···················· | ···················· | ···················· | |
| D | 6154 hCoV-19/Bangladesh/BCSIR-NILMRC-080/2020\|EPI_ISL_4 | ···················· | ···················· | ···················· | ···················· | ···················· | ···················· | |
| D | 6155 hCoV-19/Bangladesh/BCSIR-NILMRC-064/2020\|EPI_ISL_4 | ···················· | ···················· | ···················· | ···················· | ···················· | ···················· | |

| | | | | | | | | |
|---|---|---|---|---|---|---|---|---|
| D | 6156 hCoV-19/Bangladesh/BCSIR-NILMRC-066/2020\|EPI_ISL_4 | ···················· | ···················· | ···················· | ···················· | ···················· | ···················· | |
| D | 6157 hCoV-19/Bangladesh/BCSIR-NILMRC-060/2020\|EPI_ISL_4 | ···················· | ···················· | ···················· | ···················· | ···················· | ···················· | |
| D | 6158 hCoV-19/Bangladesh/BCSIR-NILMRC-062/2020\|EPI_ISL_4 | ···················· | ···················· | ···················· | ···················· | ···················· | ···················· | |
| D | 6159 hCoV-19/USA/VI-CDC-3638/2020\|EPI_ISL_464167\|2020-0 | ···················· | ···················· | ···················· | ···················· | ···················· | ···················· | |
| D | 6160 hCoV-19/England/20072030304/2020\|EPI_ISL_464168\|20 | ···················· | ···················· | ···················· | ···················· | ···················· | ···················· | |
| D | 6161 hCoV-19/England/20078003804/2020\|EPI_ISL_464169\|20 | ···················· | ···················· | ···················· | ···················· | ···················· | ···················· | |
| D | 6162 hCoV-19/England/20080000104/2020\|EPI_ISL_464175\|20 | ···················· | ···················· | ···················· | ···················· | ···················· | ···················· | |
| D | 6163 hCoV-19/England/20084031204/2020\|EPI_ISL_464178\|20 | ···················· | ···················· | ···················· | ···················· | ····R··············· | ···················· | |
| D | 6164 hCoV-19/England/20174049604/2020\|EPI_ISL_465404\|20 | ···················· | ···················· | ···················· | ···················· | ···················· | ···················· | |
| D | 6165 hCoV-19/England/20174050304/2020\|EPI_ISL_465406\|20 | ···················· | ···················· | ···················· | ···················· | ···················· | ···················· | |
| D | 6166 hCoV-19/England/20174050404/2020\|EPI_ISL_465407\|20 | ···················· | ···················· | ···················· | ···················· | ···················· | ···················· | |
| D | 6167 hCoV-19/England/20176041104/2020\|EPI_ISL_465411\|20 | ···················· | ···················· | ···················· | ···················· | ···················· | ···················· | |
| D | 6168 hCoV-19/England/20176041404/2020\|EPI_ISL_465412\|20 | ···················· | ····A··············· | ···················· | ···················· | ···················· | ···················· | |
| D | 6169 hCoV-19/England/20176047004/2020\|EPI_ISL_465417\|20 | ···················· | ···················· | ···················· | ···················· | ···················· | ···················· | |
| D | 6171 hCoV-19/England/20180025604/2020\|EPI_ISL_465430\|20 | ···················· | ···················· | ···················· | ···················· | ···················· | ···················· | |
| D | 6172 hCoV-19/England/20180026004/2020\|EPI_ISL_465433\|20 | ···················· | ···················· | ···················· | ···················· | ···················· | ···················· | |
| D | 6173 hCoV-19/England/20186075004/2020\|EPI_ISL_465471\|20 | ···················· | ···················· | ···················· | ···················· | ···················· | ···················· | |
| D | 6175 hCoV-19/England/20186077504/2020\|EPI_ISL_465475\|20 | ···················· | ···················· | ···················· | ···················· | ···················· | ···················· | |
| D | 6176 hCoV-19/England/20186077604/2020\|EPI_ISL_465476\|20 | ···················· | ···················· | ···················· | ···················· | ···················· | ···················· | |
| D | 6177 hCoV-19/England/20186078004/2020\|EPI_ISL_465477\|20 | ···················· | ···················· | ···················· | ···················· | ···················· | ·················N·N | 1 |
| D | 6180 hCoV-19/England/20186123804/2020\|EPI_ISL_465488\|20 | ···················· | ···················· | ···················· | ···················· | ···················· | ···················· | |
| D | 6181 hCoV-19/England/20186134604/2020\|EPI_ISL_465491\|20 | ···················· | ···················· | ···················· | ···················· | ···················· | ···················· | |
| D | 6183 hCoV-19/England/20188017704/2020\|EPI_ISL_465493\|20 | ···················· | ···················· | ···················· | ···················· | ···················· | ···················· | |
| D | 6184 hCoV-19/England/20188018004/2020\|EPI_ISL_465494\|20 | ···················· | ···················· | ···················· | ···················· | ···················· | ···················· | |
| D | 6185 hCoV-19/England/20188018104/2020\|EPI_ISL_465495\|20 | ···················· | ···················· | ···················· | ···················· | ···················· | ···················· | |
| D | 6186 hCoV-19/England/20188020204/2020\|EPI_ISL_465498\|20 | ···················· | ···················· | ···················· | ···················· | ···················· | ···················· | |
| D | 6187 hCoV-19/England/20188020404/2020\|EPI_ISL_465499\|20 | ···················· | ···················· | ···················· | ···················· | ···················· | ···················· | |
| D | 6188 hCoV-19/Canada/BC_00713032/2020\|EPI_ISL_466700\|202 | ···················· | ···················· | ···················· | ···················· | ···················· | ···················· | |
| D | 6189 hCoV-19/Canada/BC_00714140/2020\|EPI_ISL_466701\|202 | ···················· | ···················· | ···················· | ···················· | ···················· | ···················· | |
| D | 6190 hCoV-19/Canada/BC_00809458/2020\|EPI_ISL_466702\|202 | ···················· | ···················· | ···················· | ···················· | ···················· | ···················· | |
| D | 6191 hCoV-19/Canada/BC_01210684/2020\|EPI_ISL_466703\|202 | ···················· | ···················· | ···················· | ···················· | ···················· | ···················· | |
| D | 6192 hCoV-19/Canada/BC_01218976/2020\|EPI_ISL_466704\|202 | ···················· | ···················· | ···················· | ···················· | ···················· | ···················· | |
| D | 6193 hCoV-19/Canada/BC_01222955/2020\|EPI_ISL_466705\|202 | ···················· | ···················· | ···················· | ···················· | ···················· | ···················· | |
| D | 6194 hCoV-19/Canada/BC_01254071/2020\|EPI_ISL_466706\|202 | ···················· | ···················· | ···················· | ···················· | ···················· | ···················· | |
| D | 6195 hCoV-19/Canada/BC_01294861/2020\|EPI_ISL_466707\|202 | ···················· | ···················· | ···················· | ···················· | ···················· | ···················· | |
| D | 6196 hCoV-19/Canada/BC_01296812/2020\|EPI_ISL_466708\|202 | ···················· | ···················· | ···················· | ···················· | ···················· | ···················· | |
| D | 6197 hCoV-19/Canada/BC_01468719/2020\|EPI_ISL_466709\|202 | ···················· | ···················· | ···················· | ···················· | ···················· | ···················· | |
| D | 6198 hCoV-19/Canada/BC_01560118/2020\|EPI_ISL_466710\|202 | ···················· | ···················· | ···················· | ···················· | ···················· | ···················· | |
| D | 6199 hCoV-19/Canada/BC_01594615/2020\|EPI_ISL_466711\|202 | ···················· | ···················· | ···················· | ···················· | ···················· | ···················· | |
| D | 6200 hCoV-19/Canada/BC_02045287/2020\|EPI_ISL_466712\|202 | ···················· | ···················· | ···················· | ···················· | ···················· | ···················· | |
| D | 6201 hCoV-19/Canada/BC_02051350/2020\|EPI_ISL_466713\|202 | ···················· | ···················· | ···················· | ···················· | ···················· | ···················· | |
| D | 6202 hCoV-19/Canada/BC_02077392/2020\|EPI_ISL_466714\|202 | ···················· | ···················· | ···················· | ···················· | ···················· | ···················· | |
| D | 6203 hCoV-19/Canada/BC_02083468/2020\|EPI_ISL_466715\|202 | ···················· | ···················· | ···················· | ···················· | ···················· | ···················· | |
| D | 6204 hCoV-19/Canada/BC_02091552/2020\|EPI_ISL_466716\|202 | ···················· | ···················· | ···················· | ···················· | ···················· | ···················· | |
| D | 6205 hCoV-19/Canada/BC_02168448/2020\|EPI_ISL_466717\|202 | ···················· | ···················· | ···················· | ···················· | ···················· | ···················· | |
| D | 6206 hCoV-19/Canada/BC_02169597/2020\|EPI_ISL_466718\|202 | ···················· | ···················· | ···················· | ···················· | ···················· | ···················· | |
| D | 6207 hCoV-19/Canada/BC_02185653/2020\|EPI_ISL_466719\|202 | ···················· | ···················· | ···················· | ···················· | ···················· | ···················· | |
| D | 6208 hCoV-19/Canada/BC_02246883/2020\|EPI_ISL_466720\|202 | ···················· | ···················· | ···················· | ···················· | ···················· | ···················· | |
| D | 6209 hCoV-19/Canada/BC_02415563/2020\|EPI_ISL_466721\|202 | ···················· | ·A·················· | ···················· | ···················· | ···················· | ···················· | |
| D | 6210 hCoV-19/Canada/BC_02472119/2020\|EPI_ISL_466722\|202 | ···················· | ···················· | ···················· | ···················· | ···················· | ···················· | |
| D | 6211 hCoV-19/Canada/BC_02502338/2020\|EPI_ISL_466723\|202 | ···················· | ···················· | ···················· | ···················· | ···················· | ···················· | |
| D | 6212 hCoV-19/Canada/BC_02511020/2020\|EPI_ISL_466724\|202 | ···················· | ···················· | ···················· | ···················· | ···················· | ···················· | |
| D | 6213 hCoV-19/Canada/BC_02585169/2020\|EPI_ISL_466725\|202 | ···················· | ···················· | ···················· | ···················· | ···················· | ···················· | |
| D | 6214 hCoV-19/Canada/BC_02594336/2020\|EPI_ISL_466726\|202 | ···················· | ···················· | ···················· | ···················· | ···················· | ···················· | |
| D | 6215 hCoV-19/Canada/BC_02803451/2020\|EPI_ISL_466727\|202 | ···················· | ···················· | ···················· | ···················· | ···················· | ···················· | |
| D | 6216 hCoV-19/Canada/BC_02807114/2020\|EPI_ISL_466728\|202 | ···················· | ···················· | ···················· | ···················· | ···················· | ···················· | |
| D | 6217 hCoV-19/Canada/BC_02811162/2020\|EPI_ISL_466729\|202 | ···················· | ···················· | ···················· | ···················· | ···················· | ···················· | |
| D | 6218 hCoV-19/Canada/BC_02820264/2020\|EPI_ISL_466730\|202 | ···················· | ···················· | ···················· | ···················· | ···················· | ···················· | |
| D | 6219 hCoV-19/Canada/BC_02822314/2020\|EPI_ISL_466731\|202 | ···················· | ···················· | ···················· | ···················· | ···················· | ···················· | |
| D | 6220 hCoV-19/Canada/BC_02887476/2020\|EPI_ISL_466732\|202 | ···················· | ···················· | ···················· | ···················· | ···················· | ···················· | |
| D | 6221 hCoV-19/Canada/BC_02888326/2020\|EPI_ISL_466733\|202 | ···················· | ···················· | ···················· | ···················· | ···················· | ···················· | |
| D | 6222 hCoV-19/Canada/BC_03055797/2020\|EPI_ISL_466734\|202 | ···················· | ···················· | ···················· | ···················· | ···················· | ···················· | |
| D | 6223 hCoV-19/Canada/BC_03069740/2020\|EPI_ISL_466735\|202 | ···················· | ···················· | ···················· | ···················· | ···················· | ···················· | |
| D | 6224 hCoV-19/Canada/BC_03120111/2020\|EPI_ISL_466736\|202 | ···················· | ···················· | ···················· | ···················· | ···················· | ···················· | |
| D | 6225 hCoV-19/Canada/BC_03385157/2020\|EPI_ISL_466737\|202 | ···················· | ···················· | ···················· | ···················· | ···················· | ···················· | |
| D | 6226 hCoV-19/Canada/BC_03386598/2020\|EPI_ISL_466738\|202 | ···················· | ···················· | ···················· | ···················· | ···················· | ···················· | |
| D | 6227 hCoV-19/Canada/BC_03405412/2020\|EPI_ISL_466739\|202 | ···················· | ···················· | ···················· | ···················· | ···················· | ···················· | |
| D | 6228 hCoV-19/Canada/BC_03423088/2020\|EPI_ISL_466740\|202 | ···················· | ···················· | ···················· | ···················· | ···················· | ···················· | |
| D | 6229 hCoV-19/Canada/BC_03452391/2020\|EPI_ISL_466741\|202 | ···················· | ···················· | ···················· | ···················· | ···················· | ···················· | |
| D | 6230 hCoV-19/Canada/BC_03469645/2020\|EPI_ISL_466742\|202 | ···················· | ···················· | ···················· | ···················· | ···················· | ···················· | |
| D | 6231 hCoV-19/Canada/BC_03469815/2020\|EPI_ISL_466743\|202 | ···················· | ···················· | ···················· | ···················· | ···················· | ···················· | |
| D | 6232 hCoV-19/Canada/BC_03470015/2020\|EPI_ISL_466745\|202 | ···················· | ···················· | ···················· | ···················· | ···················· | ···················· | |
| D | 6233 hCoV-19/Canada/BC_03470522/2020\|EPI_ISL_466747\|202 | ···················· | ···················· | ···················· | ···················· | ···················· | ···················· | |
| D | 6234 hCoV-19/Canada/BC_03470614/2020\|EPI_ISL_466748\|202 | ···················· | ···················· | ···················· | ···················· | ···················· | ···················· | |
| D | 6235 hCoV-19/Canada/BC_03471049/2020\|EPI_ISL_466749\|202 | ···················· | ···················· | ···················· | ···················· | ···················· | ···················· | |
| D | 6236 hCoV-19/Canada/BC_03471188/2020\|EPI_ISL_466750\|202 | ···················· | ···················· | ···················· | ···················· | ···················· | ···················· | |
| D | 6237 hCoV-19/Canada/BC_03477384/2020\|EPI_ISL_466751\|202 | ···················· | ···················· | ···················· | ···················· | ···················· | ···················· | |
| D | 6238 hCoV-19/Canada/BC_03489456/2020\|EPI_ISL_466752\|202 | ···················· | ···················· | ···················· | ···················· | ···················· | ···················· | |

| | | | | | | | | |
|---|---|---|---|---|---|---|---|---|
| D | 6239 hCoV-19/Canada/BC_03489776/2020\|EPI_ISL_466753\|202 | ···················· | ···················· | ···················· | ···················· | ···················· | ···················· | |
| D | 6240 hCoV-19/Canada/BC_03493786/2020\|EPI_ISL_466754\|202 | ···················· | ···················· | ···················· | ···················· | ···················· | ···················· | |
| D | 6241 hCoV-19/Canada/BC_03907778/2020\|EPI_ISL_466755\|202 | ···················· | ···················· | ···················· | ···················· | ···················· | ···················· | |
| D | 6242 hCoV-19/Canada/BC_03914944/2020\|EPI_ISL_466756\|202 | ···················· | ···················· | ···················· | ···················· | ···················· | ···················· | |
| D | 6243 hCoV-19/Canada/BC_03917951/2020\|EPI_ISL_466757\|202 | ···················· | ···················· | ···················· | ···················· | ···················· | ···················· | |
| D | 6244 hCoV-19/Canada/BC_03959087/2020\|EPI_ISL_466758\|202 | ···················· | ···················· | ···················· | ···················· | ···················· | ···················· | |
| D | 6245 hCoV-19/Canada/BC_03970582/2020\|EPI_ISL_466759\|202 | ···················· | ···················· | ···················· | ···················· | ···················· | ···················· | |
| D | 6246 hCoV-19/Canada/BC_04013424/2020\|EPI_ISL_466760\|202 | ···················· | ···················· | ···················· | ···················· | ···················· | ···················· | |
| D | 6247 hCoV-19/Canada/BC_04020783/2020\|EPI_ISL_466761\|202 | ···················· | ···················· | ···················· | ···················· | ···················· | ···················· | |
| D | 6248 hCoV-19/Canada/BC_04033837/2020\|EPI_ISL_466762\|202 | ···················· | ···················· | ···················· | ···················· | ···················· | ···················· | |
| D | 6249 hCoV-19/Canada/BC_04048819/2020\|EPI_ISL_466763\|202 | ···················· | ···················· | ···················· | ···················· | ···················· | ···················· | |
| D | 6250 hCoV-19/Canada/BC_04164497/2020\|EPI_ISL_466764\|202 | ···················· | ···················· | ···················· | ···················· | ···················· | ···················· | |
| D | 6251 hCoV-19/Canada/BC_04869329/2020\|EPI_ISL_466769\|202 | ···················· | ···················· | ···················· | ···················· | ···················· | ···················· | |
| D | 6252 hCoV-19/Canada/BC_06364232/2020\|EPI_ISL_466798\|202 | ···················· | ···················· | ···················· | ···················· | ···················· | ···················· | |
| D | 6253 hCoV-19/USA/WI-WSLH-200055/2020\|EPI_ISL_471220\|202 | ···················· | ···················· | ···················· | ···················· | ···················· | ···················· | |
| D | 6254 hCoV-19/USA/WI-WSLH-200056/2020\|EPI_ISL_471221\|202 | ···················· | ···················· | ···················· | ···················· | ···················· | ···················· | |
| D | 6255 hCoV-19/USA/WI-WSLH-200059/2020\|EPI_ISL_471224\|202 | ···················· | ···················· | ···················· | ···················· | ···················· | ···················· | |
| D | 6256 hCoV-19/USA/WI-WSLH-200060/2020\|EPI_ISL_471225\|202 | ···················· | ···················· | ···················· | ···················· | ···················· | ···················· | |
| D | 6257 hCoV-19/USA/WI-WSLH-200062/2020\|EPI_ISL_471227\|202 | ···················· | ···················· | ···················· | ···················· | ···················· | ···················· | |
| D | 6258 hCoV-19/USA/WI-WSLH-200064/2020\|EPI_ISL_471229\|202 | ···················· | ···················· | ···················· | ···················· | ···················· | ···················· | |
| D | 6259 hCoV-19/USA/WI-WSLH-200065/2020\|EPI_ISL_471230\|202 | ···················· | ···················· | ···················· | ···················· | ···················· | ···················· | |
| D | 6260 hCoV-19/USA/WI-WSLH-200066/2020\|EPI_ISL_471231\|202 | ···················· | ···················· | ···················· | ············A······· | ···················· | ···················· | |
| D | 6261 hCoV-19/USA/WI-WSLH-200070/2020\|EPI_ISL_471235\|202 | ···················· | ···················· | ···················· | ···················· | ···················· | ···················· | |
| D | 6262 hCoV-19/USA/WI-WSLH-200075/2020\|EPI_ISL_471240\|202 | ···················· | ···················· | ···················· | ···················· | ···················· | ···················· | |
| D | 6263 hCoV-19/USA/WI-WSLH-200076/2020\|EPI_ISL_471241\|202 | ···················· | ···················· | ···················· | ···················· | ···················· | ···················· | |
| D | 6264 hCoV-19/USA/WI-WSLH-200077/2020\|EPI_ISL_471242\|202 | ···················· | ···················· | ···················· | ···················· | ···················· | ···················· | |
| D | 6265 hCoV-19/USA/WI-WSLH-200079/2020\|EPI_ISL_471244\|202 | ···················· | ···················· | ···················· | ···················· | ···················· | ···················· | |
| D | 6266 hCoV-19/USA/WI-WSLH-200081/2020\|EPI_ISL_471245\|202 | ···················· | ···················· | ···················· | ···················· | ···················· | ···················· | |
| D | 6267 hCoV-19/USA/WI-WSLH-200082/2020\|EPI_ISL_471246\|202 | ···················· | ···················· | ···················· | ···················· | ···················· | ···················· | |
| D | 6268 hCoV-19/USA/WI-WSLH-200083/2020\|EPI_ISL_471247\|202 | ···················· | ···················· | ···················· | ···················· | ···················· | ···················· | |
| D | 6269 hCoV-19/USA/WI-WSLH-200084/2020\|EPI_ISL_471248\|202 | ···················· | ···················· | ···················· | ···················· | ···················· | ···················· | |
| D | 6270 hCoV-19/USA/WI-WSLH-200085/2020\|EPI_ISL_471249\|202 | ···················· | ···················· | ···················· | ···················· | ···················· | ···················· | |
| D | 6271 hCoV-19/USA/WI-WSLH-200086/2020\|EPI_ISL_471250\|202 | ···················· | ···················· | ···················· | ···················· | ···················· | ···················· | |
| D | 6272 hCoV-19/USA/WI-WSLH-200087/2020\|EPI_ISL_471251\|202 | ···················· | ···················· | ···················· | ···················· | ···················· | ···················· | |
| D | 6273 hCoV-19/USA/WI-WSLH-200088/2020\|EPI_ISL_471252\|202 | ···················· | ···················· | ···················· | ···················· | ···················· | ···················· | |
| D | 6274 hCoV-19/USA/WI-WSLH-200089/2020\|EPI_ISL_471253\|202 | ···················· | ···················· | ···················· | ···················· | ···················· | ···················· | |
| D | 6275 hCoV-19/USA/WI-WSLH-200090/2020\|EPI_ISL_471254\|202 | ···················· | ···················· | ···················· | ···················· | ···················· | ···················· | |
| D | 6276 hCoV-19/USA/WI-WSLH-200091/2020\|EPI_ISL_471255\|202 | ···················· | ···················· | ···················· | ···················· | ···················· | ···················· | |
| D | 6277 hCoV-19/USA/WI-WSLH-200092/2020\|EPI_ISL_471256\|202 | ···················· | ···················· | ···················· | ···················· | ···················· | ···················· | |
| D | 6278 hCoV-19/USA/WI-WSLH-200094/2020\|EPI_ISL_471258\|202 | ···················· | ···················· | ···················· | ···················· | ···················· | ···················· | |
| D | 6279 hCoV-19/USA/WI-WSLH-200096/2020\|EPI_ISL_471260\|202 | ···················· | ···················· | ···················· | ···················· | ···················· | ···················· | |
| D | 6280 hCoV-19/USA/WI-WSLH-200097/2020\|EPI_ISL_471261\|202 | ···················· | ···················· | ···················· | ···················· | ···················· | ···················· | |
| D | 6281 hCoV-19/USA/WI-WSLH-200098/2020\|EPI_ISL_471262\|202 | ···················· | ···················· | ···················· | ···················· | ···················· | ···················· | |
| D | 6282 hCoV-19/USA/WI-WSLH-200100/2020\|EPI_ISL_471264\|202 | ···················· | ···················· | ···················· | ···················· | ···················· | ···················· | |
| D | 6283 hCoV-19/USA/WI-WSLH-200101/2020\|EPI_ISL_471265\|202 | ···················· | ···················· | ···················· | ···················· | ···················· | ···················· | |
| D | 6284 hCoV-19/USA/WI-WSLH-200102/2020\|EPI_ISL_471266\|202 | ···················· | ···················· | ···················· | ···················· | ···················· | ···················· | |
| D | 6285 hCoV-19/Ecuador/USFQ-020/2020\|EPI_ISL_471267\|2020- | ···················· | ···················· | ···················· | ···················· | ···················· | ···················· | |
| D | 6286 hCoV-19/Ecuador/USFQ-045/2020\|EPI_ISL_471268\|2020- | ···················· | ···················· | ···················· | ···················· | ···················· | ···················· | |
| D | 6287 hCoV-19/Ecuador/USFQ-707/2020\|EPI_ISL_471269\|2020- | ···················· | ···················· | ···················· | ···················· | ···················· | ···················· | |
| D | 6288 hCoV-19/Ecuador/USFQ-112/2020\|EPI_ISL_471270\|2020- | ···················· | ···················· | ···················· | ···················· | ···················· | ···················· | |
| D | 6289 hCoV-19/Ecuador/USFQ-106/2020\|EPI_ISL_471271\|2020- | ···················· | ···················· | ···················· | ···················· | ···················· | ···················· | |
| D | 6290 hCoV-19/Wales/PHWC-15C8DF/2020\|EPI_ISL_472553\|2020 | ···················· | ···················· | ···················· | ···················· | ···················· | ···················· | |
| D | 6291 hCoV-19/Wales/PHWC-15C963/2020\|EPI_ISL_472561\|2020 | ···················· | ···················· | ···················· | ···················· | ···················· | ···················· | |
| D | 6292 hCoV-19/Wales/PHWC-15C9BE/2020\|EPI_ISL_472565\|2020 | ···················· | ···················· | ···················· | ···················· | ···················· | ···················· | |
| D | 6293 hCoV-19/Wales/PHWC-15CAD9/2020\|EPI_ISL_472579\|2020 | ···················· | ···················· | ···················· | ···················· | ···················· | ···················· | |
| D | 6294 hCoV-19/Wales/PHWC-15CAF7/2020\|EPI_ISL_472580\|2020 | ···················· | ···················· | ···················· | ···················· | ···················· | ···················· | |
| D | 6295 hCoV-19/Wales/PHWC-15CB8B/2020\|EPI_ISL_472589\|2020 | ···················· | ···················· | ···················· | ···················· | ···················· | ···················· | |
| D | 6296 hCoV-19/Wales/PHWC-15CC3D/2020\|EPI_ISL_472598\|2020 | ···················· | ···················· | ···················· | ···················· | ···················· | ···················· | |
| D | 6297 hCoV-19/Scotland/EDB5720/2020\|EPI_ISL_473842\|2020- | ···················· | ···················A | ···················· | ···················· | ···················· | ···················· | 1 |
| D | 6298 hCoV-19/Scotland/EDB5722/2020\|EPI_ISL_473843\|2020- | ···················· | ···················· | ···················· | ···················· | ···················· | ···················· | |
| D | 6299 hCoV-19/Scotland/EDB5724/2020\|EPI_ISL_473845\|2020- | ···················· | ···················· | ···················· | ···················· | ···················· | ···················· | |
| D | 6300 hCoV-19/Scotland/EDB6140/2020\|EPI_ISL_473882\|2020- | ···················· | ···················· | ···················· | ···················· | ···················· | ···················· | |
| D | 6301 hCoV-19/Scotland/EDB6142/2020\|EPI_ISL_473883\|2020- | ···················· | ···················· | ···················· | ···················· | ···················· | ···················· | |
| D | 6302 hCoV-19/Scotland/EDB6143/2020\|EPI_ISL_473884\|2020- | ···················· | ···················· | ···················· | ···················· | ···················· | ···················· | |
| D | 6303 hCoV-19/Scotland/EDB6146/2020\|EPI_ISL_473885\|2020- | ···················· | ···················· | ···················· | ···················· | ···················· | ···················· | |
| D | 6304 hCoV-19/Scotland/EDB6147/2020\|EPI_ISL_473886\|2020- | ···················· | ···················A | ···················· | ···················· | ···················· | ···················· | 1 |
| D | 6305 hCoV-19/Scotland/EDB6148/2020\|EPI_ISL_473887\|2020- | ···················· | ···················A | ···················· | ···················· | ···················· | ···················· | 1 |
| D | 6306 hCoV-19/Scotland/EDB6151/2020\|EPI_ISL_473889\|2020- | ···················· | ·A·················· | ···················· | ···················· | ···················· | ···················· | |
| D | 6307 hCoV-19/Scotland/EDB6152/2020\|EPI_ISL_473890\|2020- | ···················· | ···················A | ···················· | ···················· | ···················· | ···················· | 1 |
| D | 6308 hCoV-19/Scotland/EDB6153/2020\|EPI_ISL_473891\|2020- | ···················· | ···················A | ···················· | ···················· | ···················· | ···················· | 1 |
| D | 6309 hCoV-19/Scotland/EDB6156/2020\|EPI_ISL_473892\|2020- | ···················· | ···················· | ···················· | ···················· | ···················· | ···················· | |
| D | 6310 hCoV-19/Scotland/EDB6157/2020\|EPI_ISL_473893\|2020- | ···················· | ···················· | ···················· | ···················· | ···················· | ···················· | |
| D | 6311 hCoV-19/Scotland/EDB6158/2020\|EPI_ISL_473894\|2020- | ···················· | ···················· | ···················· | ···················· | ···················· | ···················· | |
| D | 6312 hCoV-19/Scotland/EDB6160/2020\|EPI_ISL_473895\|2020- | ···················· | ···················A | ···················· | ···················· | ···················· | ···················· | 1 |
| D | 6313 hCoV-19/Scotland/EDB6164/2020\|EPI_ISL_473896\|2020- | ···················· | ···················A | ···················· | ···················· | ···················· | ···················· | 1 |
| D | 6314 hCoV-19/Scotland/EDB6165/2020\|EPI_ISL_473897\|2020- | ···················· | ···················A | ···················· | ···················· | ···················· | ···················· | 1 |
| D | 6315 hCoV-19/Scotland/EDB6167/2020\|EPI_ISL_473898\|2020- | ···················· | ···················A | ···················· | ···················· | ···················· | ···················· | 1 |
| D | 6316 hCoV-19/Scotland/EDB6168/2020\|EPI_ISL_473899\|2020- | ···················· | ···················A | ···················· | ···················· | ···················· | ···················· | 1 |

| | | | | | | | | |
|---|---|---|---|---|---|---|---|---|
| D | 6317 | hCoV-19/Switzerland/BE-ETHZ-140049/2020\|EPI_ISL_47 | ···················· | ·A·················· | ···················· | ···················· | ···················· | ···················· |
| D | 6318 | hCoV-19/Switzerland/BL-ETHZ-140050/2020\|EPI_ISL_47 | ···················· | ···················· | ···················· | ···················· | ······S············· | ···················· |
| D | 6319 | hCoV-19/Switzerland/BL-ETHZ-140051/2020\|EPI_ISL_47 | ···················· | ···················· | ···················· | ···················· | ···················· | ···················· |
| D | 6321 | hCoV-19/Switzerland/UR-ETHZ-140053/2020\|EPI_ISL_47 | ···················· | ···················· | ···················· | ···················· | ······S············· | ···················· |
| D | 6322 | hCoV-19/Switzerland/BE-ETHZ-140055/2020\|EPI_ISL_47 | ···················· | ···················· | ···················· | ···················· | ······V············· | ···················· |
| D | 6323 | hCoV-19/Switzerland/BL-ETHZ-140056/2020\|EPI_ISL_47 | ···················· | ···················· | ···················· | ···················· | ······S············· | ···················· |
| D | 6324 | hCoV-19/Switzerland/UR-ETHZ-140058/2020\|EPI_ISL_47 | ···················· | ···················· | ···················· | ···················· | ···················· | ···················· |
| D | 6325 | hCoV-19/Switzerland/BS-ETHZ-140059/2020\|EPI_ISL_47 | ···················· | ···················· | ···················· | ···················· | ······S············· | ···················· |
| D | 6326 | hCoV-19/Switzerland/BL-ETHZ-140062/2020\|EPI_ISL_47 | ···················· | ·A·················· | ···················· | ···················· | ···················· | ···················· |
| D | 6328 | hCoV-19/Switzerland/LU-ETHZ-140065/2020\|EPI_ISL_47 | ···················· | ·A·················· | ···················· | ···················· | ······S············· | ···················· |
| D | 6329 | hCoV-19/Switzerland/TG-ETHZ-140066/2020\|EPI_ISL_47 | ···················· | ···················· | ···················· | ···················· | ···················· | ···················· |
| D | 6330 | hCoV-19/Sweden/20-08356/2020\|EPI_ISL_476140\|2020-0 | ···················· | ···················· | ···················· | ···················· | ···················· | ···················· |
| D | 6331 | hCoV-19/Sweden/20-08360/2020\|EPI_ISL_476141\|2020-0 | ···················· | ···················· | ···················· | ···················· | ···················· | ···················· |
| D | 6332 | hCoV-19/Sweden/20-08533/2020\|EPI_ISL_476143\|2020-0 | ···················· | ···················· | ···················· | ···················· | ···················· | ···················· |
| D | 6333 | hCoV-19/Senegal/57065/2020\|EPI_ISL_476148\|2020-06- | ···················· | ···················· | ···················· | ···················· | ···················· | ···················· |
| D | 6334 | hCoV-19/Senegal/57476/2020\|EPI_ISL_476149\|2020-06- | ···················· | ···················· | ···················· | ···················· | ···················· | ···················· |
| D | 6335 | hCoV-19/Australia/NSW588/2020\|EPI_ISL_478700\|2020- | ···················· | ···················· | ···················· | ···················· | ···················· | ···················· |
| D | 6336 | hCoV-19/Australia/NSW589/2020\|EPI_ISL_478701\|2020- | ···················· | ···················· | ···················· | ···················· | ···················· | ···················· |
| D | 6337 | hCoV-19/Australia/NSW590/2020\|EPI_ISL_478702\|2020- | ···················· | ···················· | ···················· | ···················· | ···················· | ···················· |
| D | 6338 | hCoV-19/Australia/NSW591/2020\|EPI_ISL_478703\|2020- | ···················· | ···················· | ···················· | ···················· | ···················· | ···················· |
| D | 6339 | hCoV-19/Australia/NSW592/2020\|EPI_ISL_478704\|2020- | ···················· | ···················· | ···················· | ···················· | ···················· | ···················· |
| D | 6340 | hCoV-19/Australia/NSW593/2020\|EPI_ISL_478705\|2020- | ···················· | ···················· | ···················· | ···················· | ···················· | ···················· |
| D | 6341 | hCoV-19/Australia/NSW594/2020\|EPI_ISL_478706\|2020- | ···················· | ···················· | ···················· | ···················· | ···················· | ···················· |
| D | 6342 | hCoV-19/Australia/NSW595/2020\|EPI_ISL_478707\|2020- | ···················· | ···················· | ···················· | ···················· | ···················· | ···················· |
| D | 6343 | hCoV-19/Australia/NSW596/2020\|EPI_ISL_478708\|2020- | ···················· | ···················· | ···················· | ···················· | ···················· | ···················· |
| D | 6344 | hCoV-19/Australia/NSW597/2020\|EPI_ISL_478709\|2020- | ···················· | ···················· | ···················· | ···················· | ···················· | ···················· |
| D | 6345 | hCoV-19/Australia/NSW598/2020\|EPI_ISL_478710\|2020- | ···················· | ···················· | ···················· | ···················· | ···················· | ···················· |
| D | 6346 | hCoV-19/Australia/NSW599/2020\|EPI_ISL_478711\|2020- | ···················· | ···················· | ···················· | ···················· | ···················· | ···················· |
| D | 6347 | hCoV-19/Australia/NSW600/2020\|EPI_ISL_478712\|2020- | ···················· | ···················· | ···················· | ···················· | ···················· | ···················· |
| D | 6348 | hCoV-19/Australia/NSW601/2020\|EPI_ISL_478713\|2020- | ···················· | ···················· | ···················· | ···················· | ···················· | ···················· |
| D | 6349 | hCoV-19/Australia/NSW602/2020\|EPI_ISL_478714\|2020- | ···················· | ···················· | ···················· | ···················· | ···················· | ···················· |
| D | 6350 | hCoV-19/Australia/NSW603/2020\|EPI_ISL_478715\|2020- | ···················· | ···················· | ···················· | ···················· | ···················· | ···················· |
| D | 6351 | hCoV-19/Australia/NSW604/2020\|EPI_ISL_478716\|2020- | ···················· | ···················· | ···················· | ···················· | ···················· | ···················· |
| D | 6352 | hCoV-19/Australia/NSW605/2020\|EPI_ISL_478717\|2020- | ···················· | ···················· | ···················· | ···················· | ···················· | ···················· |
| D | 6353 | hCoV-19/Australia/NSW606/2020\|EPI_ISL_478718\|2020- | ···················· | ···················· | ···················· | ···················· | ···················· | ···················· |
| D | 6354 | hCoV-19/England/NORW-EA52B/2020\|EPI_ISL_478719\|202 | ···················· | ···················· | ···················· | ···················· | ···················· | ···················· |
| D | 6356 | hCoV-19/England/NOTT-112037/2020\|EPI_ISL_478721\|20 | ···················· | ···················· | ···················· | ···················· | ···················· | ···················· |
| D | 6357 | hCoV-19/England/NOTT-112046/2020\|EPI_ISL_478722\|20 | ···················· | ···················· | ···················· | ···················· | ···················· | ···················· |
| D | 6358 | hCoV-19/England/NOTT-112064/2020\|EPI_ISL_478724\|20 | ···················· | ···················· | ···················· | ···················· | ···················· | ···················· |
| D | 6359 | hCoV-19/England/NOTT-112091/2020\|EPI_ISL_478725\|20 | ···················· | ···················· | ···················· | ···················· | ···················· | ···················· |
| D | 6360 | hCoV-19/England/OXON-AF337/2020\|EPI_ISL_478729\|202 | ···················· | ···················· | ···················· | ···················· | ···················· | ···················· |
| D | 6361 | hCoV-19/England/OXON-AF364/2020\|EPI_ISL_478730\|202 | ···················· | ···················· | ···················· | ···················· | ···················· | ···················· |
| D | 6362 | hCoV-19/England/OXON-AF3BF/2020\|EPI_ISL_478731\|202 | ···················· | ···················· | ···················· | ···················· | ···················· | ···················· |
| D | 6363 | hCoV-19/England/OXON-AF3DD/2020\|EPI_ISL_478732\|202 | ···················· | ···················· | ···················· | ···················· | ···················· | ···················· |
| D | 6364 | hCoV-19/England/OXON-AF3EC/2020\|EPI_ISL_478733\|202 | ···················· | ···················· | ···················· | ···················· | ···················· | ···················· |
| D | 6365 | hCoV-19/England/OXON-AF3FB/2020\|EPI_ISL_478734\|202 | ···················· | ···················· | ···················· | ···················· | ···················· | ···················· |
| D | 6366 | hCoV-19/England/OXON-AF407/2020\|EPI_ISL_478735\|202 | ···················· | ···················· | ···················· | ···················· | ···················· | ···················· |
| D | 6367 | hCoV-19/England/OXON-AF416/2020\|EPI_ISL_478736\|202 | ···················· | ···················· | ···················· | ···················· | ···················· | ···················· |
| D | 6368 | hCoV-19/England/OXON-AF425/2020\|EPI_ISL_478737\|202 | ···················· | ···················· | ···················· | ···················· | ···················· | ···················· |
| D | 6369 | hCoV-19/England/OXON-AF434/2020\|EPI_ISL_478738\|202 | ···················· | ···················· | ···················· | ···················· | ···················· | ···················· |
| D | 6370 | hCoV-19/England/OXON-AF443/2020\|EPI_ISL_478739\|202 | ···················· | ···················· | ···················· | ···················· | ···················· | ···················· |
| D | 6371 | hCoV-19/England/OXON-AF452/2020\|EPI_ISL_478740\|202 | ···················· | ···················· | ···················· | ···················· | ···················· | ···················· |
| D | 6372 | hCoV-19/England/OXON-AF48F/2020\|EPI_ISL_478742\|202 | ···················· | ···················· | ···················· | ···················· | ···················· | ···················· |
| D | 6373 | hCoV-19/England/OXON-AF4CB/2020\|EPI_ISL_478743\|202 | ···················· | ···················· | ···················· | ···················· | ···················· | ···················· |
| D | 6374 | hCoV-19/England/OXON-AF4DA/2020\|EPI_ISL_478744\|202 | ···················· | ···················· | ···················· | ···················· | ···················· | ···················· |
| D | 6375 | hCoV-19/England/OXON-AF4E9/2020\|EPI_ISL_478745\|202 | ···················· | ···················· | ···················· | ···················· | ···················· | ···················· |
| D | 6376 | hCoV-19/England/OXON-AF4F8/2020\|EPI_ISL_478746\|202 | ···················· | ···················· | ···················· | ···················· | ···················· | ···················· |
| D | 6377 | hCoV-19/England/OXON-AF513/2020\|EPI_ISL_478747\|202 | ···················· | ···················· | ···················· | ···················· | ···················· | ···················· |
| D | 6378 | hCoV-19/England/OXON-AF531/2020\|EPI_ISL_478748\|202 | ···················· | ···················· | ···················· | ···················· | ···················· | ···················· |
| D | 6379 | hCoV-19/England/OXON-AF58C/2020\|EPI_ISL_478752\|202 | ···················· | ···················· | ···················· | ···················· | ···················· | ···················· |
| D | 6380 | hCoV-19/England/OXON-AF5C8/2020\|EPI_ISL_478753\|202 | ···················· | ···················· | ···················· | ···················· | ···················· | ···················· |
| D | 6381 | hCoV-19/England/OXON-AF5D7/2020\|EPI_ISL_478754\|202 | ···················· | ···················· | ···················· | ···················· | ···················· | ···················· |
| D | 6382 | hCoV-19/England/OXON-AF5E6/2020\|EPI_ISL_478755\|202 | ···················· | ···················· | ···················· | ···················· | ···················· | ···················· |
| D | 6383 | hCoV-19/England/OXON-AF601/2020\|EPI_ISL_478756\|202 | ···················· | ···················· | ···················· | ···················· | ···················· | ···················· |
| D | 6384 | hCoV-19/England/OXON-AF610/2020\|EPI_ISL_478757\|202 | ···················· | ···················· | ···················· | ···················· | ···················· | ···················· |
| D | 6385 | hCoV-19/England/OXON-AF64D/2020\|EPI_ISL_478758\|202 | ···················· | ···················· | ···················· | ···················· | ···················· | ···················· |
| D | 6386 | hCoV-19/England/OXON-AF65C/2020\|EPI_ISL_478759\|202 | ···················· | ···················· | ···················· | ···················· | ···················· | ···················· |
| D | 6387 | hCoV-19/England/OXON-AF66B/2020\|EPI_ISL_478760\|202 | ···················· | ···················· | ···················· | ···················· | ···················· | ···················· |
| D | 6388 | hCoV-19/England/OXON-AF67A/2020\|EPI_ISL_478761\|202 | ···················· | ···················· | ···················· | ···················· | ···················· | ···················· |
| D | 6389 | hCoV-19/England/OXON-AF6A7/2020\|EPI_ISL_478762\|202 | ···················· | ···················· | ···················· | ···················· | ···················· | ···················· |
| D | 6390 | hCoV-19/England/OXON-AF6C5/2020\|EPI_ISL_478763\|202 | ···················· | ·A·················· | ···················· | ···················· | ···················· | ···················· |
| D | 6391 | hCoV-19/England/OXON-AF70E/2020\|EPI_ISL_478764\|202 | ···················· | ···················· | ···················· | ···················· | ···················· | ···················· |
| D | 6392 | hCoV-19/England/OXON-AF72C/2020\|EPI_ISL_478765\|202 | ···················· | ···················· | ···················· | ···················· | ···················· | ···················· |
| D | 6393 | hCoV-19/England/OXON-AF759/2020\|EPI_ISL_478768\|202 | ···················· | ···················· | ···················· | ···················· | ···················· | ···················· |
| D | 6394 | hCoV-19/England/OXON-AF777/2020\|EPI_ISL_478769\|202 | ···················· | ···················· | ···················· | ···················· | ···················· | ···················· |
| D | 6395 | hCoV-19/England/OXON-AF7B3/2020\|EPI_ISL_478771\|202 | ···················· | ···················· | ···················· | ···················· | ···················· | ···················· |
| D | 6396 | hCoV-19/England/OXON-AF7C2/2020\|EPI_ISL_478772\|202 | ···················· | ···················· | ···················· | ···················· | ···················· | ···················· |
| D | 6397 | hCoV-19/England/OXON-AF7D1/2020\|EPI_ISL_478773\|202 | ···················· | ···················· | ···················· | ···················· | ···················· | ···················· |

| | | | | | | | |
|---|---|---|---|---|---|---|---|
| D | 6398 hCoV-19/England/OXON-AF7FF/2020\|EPI_ISL_478774\|202 | ···················· | ···················· | ···················· | ···················· | ···················· | ···················· |
| D | 6399 hCoV-19/England/OXON-AF81A/2020\|EPI_ISL_478775\|202 | ···················· | ···················· | ···················· | ···················· | ···················· | ···················· |
| D | 6400 hCoV-19/England/OXON-AF838/2020\|EPI_ISL_478776\|202 | ···················· | ···················· | ···················· | ···················· | ···················· | ···················· |
| D | 6401 hCoV-19/Canada/ON-UHTC_0056/2020\|EPI_ISL_464059\|20 | ···················· | ···················· | ···················· | ···················· | ···················· | ···················· |
| D | 6402 hCoV-19/Canada/ON-UHTC_0045/2020\|EPI_ISL_464061\|20 | ···················· | ···················· | ···················· | ···················· | ···················· | ···················· |
| D | 6403 hCoV-19/Canada/ON-UHTC_0074/2020\|EPI_ISL_464062\|20 | ···················· | ···················· | ···················· | ···················· | ···················· | ···················· |
| D | 6404 hCoV-19/Canada/ON-UHTC_0051/2020\|EPI_ISL_464063\|20 | ···················· | ···················· | ···················· | ···················· | ···················· | ···················· |
| D | 6405 hCoV-19/Canada/ON-UHTC_0081/2020\|EPI_ISL_464064\|20 | ···················· | ···················· | ···················· | ···················· | ···················· | ···················· |
| D | 6406 hCoV-19/Belgium/BT-0407271/2020\|EPI_ISL_464065\|202 | ···················· | ···················· | ···················· | ···················· | ···················· | ···················· |
| D | 6407 hCoV-19/Belgium/GSS-0407272/2020\|EPI_ISL_464066\|20 | ···················· | ···················· | ···················· | ···················· | ···················· | ···················· |
| D | 6408 hCoV-19/Belgium/MA-0406273/2020\|EPI_ISL_464067\|202 | ···················· | ···················· | ···················· | ···················· | ···················· | ···················· |
| D | 6409 hCoV-19/Belgium/MI-0407274/2020\|EPI_ISL_464068\|202 | ···················· | ···················· | ···················· | ···················· | ···················· | ···················· |
| D | 6410 hCoV-19/Belgium/WLM-0406275/2020\|EPI_ISL_464069\|20 | ···················· | ···················· | ···················· | ···················· | ···················· | ···················· |
| D | 6411 hCoV-19/Belgium/CK-0404276/2020\|EPI_ISL_464070\|202 | ···················· | ···················· | ···················· | ···················· | ···················· | ···················· |
| D | 6412 hCoV-19/Belgium/TVMT-0404277/2020\|EPI_ISL_464071\|2 | ···················· | ···················· | ···················· | ···················· | ···················· | ···················· |
| D | 6413 hCoV-19/Belgium/LC-0406278/2020\|EPI_ISL_464072\|202 | ···················· | ···················· | ···················· | ···················· | ···················· | ···················· |
| D | 6414 hCoV-19/Belgium/VSL-0407279/2020\|EPI_ISL_464073\|20 | ···················· | ···················· | ···················· | ···················· | ···················· | ···················· |
| D | 6415 hCoV-19/Belgium/BD-0407280/2020\|EPI_ISL_464074\|202 | ···················· | ···················· | ···················· | ···················· | ···················· | ···················· |
| D | 6416 hCoV-19/Belgium/rega-0408281/2020\|EPI_ISL_464075\|2 | ···················· | ···················· | ···················· | ···················· | ···················· | ···················· |
| D | 6417 hCoV-19/Belgium/rega-0407282/2020\|EPI_ISL_464076\|2 | ···················· | ···················· | ···················· | ···················· | ···················· | ···················· |
| D | 6418 hCoV-19/Belgium/rega-0407283/2020\|EPI_ISL_464077\|2 | ···················· | ···················· | ···················· | ···················· | ···················· | ···················· |
| D | 6419 hCoV-19/Belgium/rega-0408284/2020\|EPI_ISL_464078\|2 | ···················· | ···················· | ···················· | ···················· | ···················· | ···················· |
| D | 6420 hCoV-19/Belgium/rega-0406285/2020\|EPI_ISL_464079\|2 | ···················· | ···················· | ···················· | ···················· | ···················· | ···················· |
| D | 6421 hCoV-19/Belgium/rega-0408286/2020\|EPI_ISL_464080\|2 | ···················· | ···················· | ···················· | ···················· | ···················· | ···················· |
| D | 6422 hCoV-19/Belgium/rega-0407287/2020\|EPI_ISL_464081\|2 | ···················· | ···················· | ···················· | ···················· | ···················· | ···················· |
| D | 6423 hCoV-19/Belgium/rega-0405288/2020\|EPI_ISL_464082\|2 | ···················· | ···················· | ···················· | ···················· | ···················· | ···················· |
| D | 6424 hCoV-19/Belgium/rega-0405289/2020\|EPI_ISL_464083\|2 | ···················· | ···················· | ···················· | ···················· | ···················· | ···················· |
| D | 6425 hCoV-19/Belgium/rega-0403290/2020\|EPI_ISL_464084\|2 | ···················· | ···················· | ···················· | ···················· | ···················· | ···················· |
| D | 6426 hCoV-19/Belgium/VJ-0409291/2020\|EPI_ISL_464085\|202 | ···················· | ···················· | ···················· | ···················· | ···················· | ···················· |
| D | 6427 hCoV-19/Belgium/RD-0408292/2020\|EPI_ISL_464086\|202 | ···················· | ···················· | ···················· | ···················· | ···················· | ···················· |
| D | 6428 hCoV-19/Belgium/HC-0408293/2020\|EPI_ISL_464087\|202 | ···················· | ···················· | ···················· | ···················· | ···················· | ···················· |
| D | 6429 hCoV-19/Belgium/LEV-0409294/2020\|EPI_ISL_464088\|20 | ···················· | ···················· | ···················· | ···················· | ···················· | ···················· |
| D | 6430 hCoV-19/Belgium/LKJ-0409295/2020\|EPI_ISL_464089\|20 | ···················· | ···················· | ···················· | ···················· | ···················· | ···················· |
| D | 6431 hCoV-19/Belgium/SA-0409296/2020\|EPI_ISL_464090\|202 | ···················· | ···················· | ···················· | ···················· | ···················· | ···················· |
| D | 6432 hCoV-19/Taiwan/CGMH-CGU-28/2020\|EPI_ISL_464092\|202 | ···················· | ···················· | ···················· | ···················· | ···················· | ···················· |
| D | 6433 hCoV-19/Taiwan/CGMH-CGU-29/2020\|EPI_ISL_464093\|202 | ···················· | ···················· | ···················· | ···················· | ···················· | ···················· |
| D | 6434 hCoV-19/Taiwan/CGMH-CGU-30/2020\|EPI_ISL_464094\|202 | ···················· | ·A·················· | ···················· | ···················· | ···················· | ···················· |
| D | 6435 hCoV-19/England/20099069004/2020\|EPI_ISL_464214\|20 | ···················· | ···················· | ···················· | ···················· | ··N················· | ···················· |
| D | 6436 hCoV-19/England/20099069504/2020\|EPI_ISL_464215\|20 | ···················· | ···················· | ···················· | ···················· | ···················· | ···················· |
| D | 6438 hCoV-19/England/20099070104/2020\|EPI_ISL_464217\|20 | ···················· | ···················· | ···················· | ···················· | ···················· | ···················· |
| D | 6439 hCoV-19/England/20099071404/2020\|EPI_ISL_464218\|20 | ···················· | ···················· | ···················· | ···················· | ···················· | ···················· |
| D | 6440 hCoV-19/England/20099071804/2020\|EPI_ISL_464219\|20 | ···················· | ···················· | ···················· | ···················· | ···················· | ···················· |
| D | 6442 hCoV-19/England/20099072204/2020\|EPI_ISL_464223\|20 | ···················· | ···················· | ···················· | ···················· | ···················· | ···················· |
| D | 6443 hCoV-19/England/20099072604/2020\|EPI_ISL_464224\|20 | ···················· | ···················· | ···················· | ···················· | ···················· | ···················· |
| D | 6444 hCoV-19/England/20099089404/2020\|EPI_ISL_464227\|20 | ···················· | ···················· | ···················· | ···················· | ···················· | ···················· |
| D | 6445 hCoV-19/England/20099089504/2020\|EPI_ISL_464228\|20 | ···················· | ···················· | ···················· | ···················· | ···················· | ···················· |
| D | 6446 hCoV-19/England/20099089604/2020\|EPI_ISL_464229\|20 | ···················· | ···················· | ···················· | ···················· | ···················· | ···················· |
| D | 6447 hCoV-19/England/20100000904/2020\|EPI_ISL_464238\|20 | ···················· | ···················· | ···················· | ···················· | ···················· | ···················· |
| D | 6448 hCoV-19/England/20100001204/2020\|EPI_ISL_464241\|20 | ···················· | ···················· | ···················· | ···················· | ···················· | ···················· |
| D | 6449 hCoV-19/England/20100004602/2020\|EPI_ISL_464248\|20 | ···················· | ···················· | ···················· | ···················· | ··N················· | ···················· |
| D | 6450 hCoV-19/England/20100005504/2020\|EPI_ISL_464252\|20 | ···················· | ···················· | ···················· | ···················· | ···················· | ···················· |
| D | 6451 hCoV-19/England/20100005604/2020\|EPI_ISL_464253\|20 | ···················· | ···················· | ···················· | ···················· | ···················· | ···················· |
| D | 6452 hCoV-19/England/20174019504/2020\|EPI_ISL_465390\|20 | ···················· | ···················· | ···················· | ···················· | ···················· | ···················· |
| D | 6453 hCoV-19/England/20174036804/2020\|EPI_ISL_465396\|20 | ···················· | ···················· | ···················· | ···················· | ···················· | ···················· |
| D | 6454 hCoV-19/England/20174037704/2020\|EPI_ISL_465397\|20 | ···················· | ···················· | ···················· | ···················· | ···················· | ···················· |
| D | 6455 hCoV-19/England/20174038204/2020\|EPI_ISL_465398\|20 | ···················· | ···················· | ···················· | ···················· | ···················· | ···················· |
| D | 6457 hCoV-19/England/20189155304/2020\|EPI_ISL_465552\|20 | ···················· | ···················· | ···················· | ···················· | ···················· | ···················· |
| D | 6458 hCoV-19/England/20189156401/2020\|EPI_ISL_465554\|20 | ···················· | ···················· | ···················· | ···················· | ···················· | ···················· |
| D | 6459 hCoV-19/England/20189156601/2020\|EPI_ISL_465555\|20 | ···················· | ···················· | ···················· | ···················· | ···················· | ···················· |
| D | 6460 hCoV-19/England/20189156801/2020\|EPI_ISL_465556\|20 | ···················· | ···················· | ···················· | ···················· | ···················· | ···················· |
| D | 6461 hCoV-19/England/20189157404/2020\|EPI_ISL_465557\|20 | ···················· | ···················· | ···················· | ···················· | ···················· | ···················· |
| D | 6462 hCoV-19/England/20189157501/2020\|EPI_ISL_465558\|20 | ···················· | ···················· | ···················· | ···················· | ···················· | ···················· |
| D | 6463 hCoV-19/England/20189158704/2020\|EPI_ISL_465559\|20 | ···················· | ···················· | ···················· | ···················· | ···················· | ···················· |
| D | 6464 hCoV-19/England/20189158801/2020\|EPI_ISL_465560\|20 | ···················· | ···················· | ···················· | ···················· | ···················· | ···················· |
| D | 6465 hCoV-19/England/20189159304/2020\|EPI_ISL_465561\|20 | ···················· | ···················· | ···················· | ···················· | ···················· | ···················· |
| D | 6466 hCoV-19/England/20189159504/2020\|EPI_ISL_465563\|20 | ···················· | ···················· | ···················· | ···················· | ···················· | ···················· |
| D | 6468 hCoV-19/England/20189159904/2020\|EPI_ISL_465565\|20 | ···················· | ···················· | ···················· | ···················· | ···················· | ···················· |
| D | 6469 hCoV-19/England/20189163501/2020\|EPI_ISL_465566\|20 | ···················· | ···················· | ···················· | ···················· | ···················· | ···················· |
| D | 6471 hCoV-19/England/20189166901/2020\|EPI_ISL_465570\|20 | ···················· | ···················· | ···················· | ···················· | ···················· | ···················· |
| D | 6472 hCoV-19/England/20189176701/2020\|EPI_ISL_465574\|20 | ···················· | ···················· | ···················· | ···················· | ···················· | ···················· |
| D | 6473 hCoV-19/England/20189177001/2020\|EPI_ISL_465576\|20 | ···················· | ···················· | ···················· | ···················· | ···················· | N··················· |
| D | 6474 hCoV-19/England/20189182004/2020\|EPI_ISL_465577\|20 | ···················· | ···················· | ···················· | ···················· | ···················· | ···················· |
| D | 6476 hCoV-19/England/20189186501/2020\|EPI_ISL_465580\|20 | ···················· | ···················· | ···················· | ···················· | ···················· | ···················· |
| D | 6477 hCoV-19/England/20190003901/2020\|EPI_ISL_465581\|20 | ···················· | ···················· | ···················· | ···················· | ···················· | ···················· |
| D | 6478 hCoV-19/England/20190007201/2020\|EPI_ISL_465582\|20 | ···················· | ···················· | ···················· | ···················· | ···················· | ···················· |
| D | 6479 hCoV-19/England/20190012401/2020\|EPI_ISL_465583\|20 | ···················· | ···················· | ···················· | ···················· | ···················· | ···················· |
| D | 6481 hCoV-19/England/20190056201/2020\|EPI_ISL_465586\|20 | ···················· | ···················· | ···················· | ···················· | ···················· | ···················· |
| D | 6482 hCoV-19/England/20190056501/2020\|EPI_ISL_465587\|20 | ···················· | ···················· | ···················· | ···················· | ···················· | ···················· |

| | | | | | | | | |
|---|---|---|---|---|---|---|---|---|
| D | 6483 hCoV-19/Germany/BY-MVP-0182/2020\|EPI_ISL_466875\|20 | .................... | .................... | .................... | .................... | .................... | .................... | |
| D | 6484 hCoV-19/Germany/BY-MVP-0184/2020\|EPI_ISL_466877\|20 | .................... | .................... | .................... | .................... | .................... | .................... | |
| D | 6485 hCoV-19/Germany/BY-MVP-0187/2020\|EPI_ISL_466880\|20 | .................... | .................... | .................... | .................... | .................... | .................... | |
| D | 6486 hCoV-19/Germany/BY-MVP-0190/2020\|EPI_ISL_466881\|20 | .................... | .................... | .................... | .................... | .................... | .................... | |
| D | 6487 hCoV-19/Germany/BY-MVP-0192/2020\|EPI_ISL_466882\|20 | .................... | .................... | .................... | .................... | .................... | .................... | |
| D | 6488 hCoV-19/Germany/BY-MVP-0193/2020\|EPI_ISL_466883\|20 | .................... | .................... | .................... | .................... | .................... | .................... | |
| D | 6489 hCoV-19/Germany/BY-MVP-0196/2020\|EPI_ISL_466884\|20 | .................... | .................... | .................... | .................... | .................... | .................... | |
| D | 6490 hCoV-19/Germany/BY-MVP-0200/2020\|EPI_ISL_466885\|20 | .................... | .................... | .................... | .................... | .................... | .................... | |
| D | 6491 hCoV-19/Germany/BY-MVP-0207/2020\|EPI_ISL_466888\|20 | .................... | .................... | .................... | .................... | .................... | .................... | |
| D | 6492 hCoV-19/Germany/BY-MVP-0208/2020\|EPI_ISL_466889\|20 | .................... | .................... | .................... | .................... | .................... | .................... | |
| D | 6493 hCoV-19/Germany/BY-MVP-0218/2020\|EPI_ISL_466891\|20 | .................... | .................... | .................... | .................... | .................... | .................... | |
| D | 6494 hCoV-19/Germany/BY-MVP-0219/2020\|EPI_ISL_466892\|20 | .................... | C................... | .................... | .................... | .................... | .................... | |
| D | 6495 hCoV-19/Germany/BY-MVP-0220/2020\|EPI_ISL_466893\|20 | .................... | .................... | .................... | .................... | .................... | .................... | |
| D | 6496 hCoV-19/Germany/BY-MVP-0225/2020\|EPI_ISL_466895\|20 | .................... | .................... | .................... | .................... | .................... | .................... | |
| D | 6497 hCoV-19/Germany/BY-MVP-0226/2020\|EPI_ISL_466896\|20 | .................... | .................... | .................... | .................... | .................... | .................... | |
| D | 6498 hCoV-19/Germany/BY-MVP-0229/2020\|EPI_ISL_466897\|20 | .................... | .................... | .................... | .................... | .................... | .................... | |
| D | 6500 hCoV-19/USA/WI-WSLH-200035/2020\|EPI_ISL_471200\|202 | .................... | .................... | .................... | .................... | .................... | .................... | |
| D | 6501 hCoV-19/USA/WI-WSLH-200036/2020\|EPI_ISL_471201\|202 | .................... | .................... | .................... | .................... | .................... | .................... | |
| D | 6502 hCoV-19/USA/WI-WSLH-200037/2020\|EPI_ISL_471202\|202 | .................... | .................... | .................... | .................... | .................... | .................... | |
| D | 6503 hCoV-19/USA/WI-WSLH-200038/2020\|EPI_ISL_471203\|202 | .................... | .................... | .................... | .................... | .................... | .................... | |
| D | 6504 hCoV-19/USA/WI-WSLH-200040/2020\|EPI_ISL_471205\|202 | .................... | .................... | .................... | .................... | .................... | .................... | |
| D | 6505 hCoV-19/USA/WI-WSLH-200041/2020\|EPI_ISL_471206\|202 | .................... | .................... | .................... | .................... | .................... | .................... | |
| D | 6506 hCoV-19/USA/WI-WSLH-200042/2020\|EPI_ISL_471207\|202 | .................... | .................... | .................... | .................... | .................... | .................... | |
| D | 6507 hCoV-19/USA/WI-WSLH-200043/2020\|EPI_ISL_471208\|202 | .................... | .................... | .................... | .................... | .................... | .................... | |
| D | 6508 hCoV-19/USA/WI-WSLH-200044/2020\|EPI_ISL_471209\|202 | .................... | .................... | .................... | .................... | .................... | .................... | |
| D | 6509 hCoV-19/USA/WI-WSLH-200046/2020\|EPI_ISL_471211\|202 | .................... | .................... | .................... | .................... | .................... | .................... | |
| D | 6510 hCoV-19/USA/WI-WSLH-200047/2020\|EPI_ISL_471212\|202 | .................... | .................... | .................... | .................... | .................... | .................... | |
| D | 6511 hCoV-19/USA/WI-WSLH-200048/2020\|EPI_ISL_471213\|202 | .................... | .................... | .................... | .................... | .................... | .................... | |
| D | 6512 hCoV-19/USA/WI-WSLH-200049/2020\|EPI_ISL_471214\|202 | .................... | .................... | .................... | .................... | .................... | .................... | |
| D | 6513 hCoV-19/USA/WI-WSLH-200050/2020\|EPI_ISL_471215\|202 | .................... | .................... | .................... | .................... | .................... | .................... | |
| D | 6514 hCoV-19/USA/WI-WSLH-200053/2020\|EPI_ISL_471218\|202 | .................... | .................... | .................... | .................... | .................... | .................... | |
| D | 6515 hCoV-19/USA/WI-WSLH-200054/2020\|EPI_ISL_471219\|202 | .................... | .................... | .................... | .................... | .................... | .................... | |
| D | 6516 hCoV-19/Wales/PHWC-15C5AB/2020\|EPI_ISL_472509\|2020 | .................... | .................... | .................... | .................... | .................... | .................... | |
| D | 6517 hCoV-19/Wales/PHWC-15C5E7/2020\|EPI_ISL_472513\|2020 | .................... | .................... | .................... | .................... | .................... | .................... | |
| D | 6518 hCoV-19/Wales/PHWC-15C611/2020\|EPI_ISL_472516\|2020 | .................... | .................... | .................... | .................... | .................... | .................... | |
| D | 6519 hCoV-19/Wales/PHWC-15C6A8/2020\|EPI_ISL_472522\|2020 | .................... | .................... | .................... | .................... | .................... | .................... | |
| D | 6520 hCoV-19/Wales/PHWC-15C6B7/2020\|EPI_ISL_472523\|2020 | .................... | ...A................ | .................... | .................... | .................... | .................... | |
| D | 6521 hCoV-19/Wales/PHWC-15C72D/2020\|EPI_ISL_472530\|2020 | .................... | .................... | .................... | .................... | .................... | .................... | |
| D | 6522 hCoV-19/Wales/PHWC-15C7B4/2020\|EPI_ISL_472538\|2020 | .................... | .................... | .................... | .................... | .................... | .................... | |
| D | 6523 hCoV-19/Wales/PHWC-15C7E1/2020\|EPI_ISL_472541\|2020 | .................... | .................... | .................... | .................... | .................... | .................... | |
| D | 6524 hCoV-19/Wales/PHWC-15C7F0/2020\|EPI_ISL_472542\|2020 | .................... | .................... | .................... | .................... | .................... | .................... | |
| D | 6525 hCoV-19/Wales/PHWC-15C82A/2020\|EPI_ISL_472545\|2020 | .................... | .................... | .................... | .................... | .................... | .................... | |
| D | 6526 hCoV-19/Wales/PHWC-15C848/2020\|EPI_ISL_472547\|2020 | .................... | .................... | .................... | .................... | .................... | .................... | |
| D | 6527 hCoV-19/Scotland/EDB4930/2020\|EPI_ISL_473800\|2020- | .................... | .................... | .................... | .................... | .................... | .................... | |
| D | 6528 hCoV-19/Scotland/EDB4948/2020\|EPI_ISL_473801\|2020- | .................... | .................... | .................... | .................... | .................... | .................... | |
| D | 6529 hCoV-19/Scotland/EDB4950/2020\|EPI_ISL_473803\|2020- | .................... | .................... | .................... | .................... | .................... | .................... | |
| D | 6530 hCoV-19/Scotland/EDB4952/2020\|EPI_ISL_473804\|2020- | .................... | .................... | .................... | .................... | .................... | .................... | |
| D | 6531 hCoV-19/Scotland/EDB4953/2020\|EPI_ISL_473805\|2020- | .................... | .................... | .................... | .................... | .................... | .................... | |
| D | 6532 hCoV-19/Scotland/EDB4956/2020\|EPI_ISL_473806\|2020- | .................... | .................... | .................... | .................... | .................... | .................... | |
| D | 6533 hCoV-19/Scotland/EDB4957/2020\|EPI_ISL_473807\|2020- | .................... | .................... | .................... | .................... | .................... | .................... | |
| D | 6534 hCoV-19/Scotland/EDB4959/2020\|EPI_ISL_473808\|2020- | .................... | .................... | .................... | .................... | .................... | .................... | |
| D | 6535 hCoV-19/Scotland/EDB4961/2020\|EPI_ISL_473809\|2020- | .................... | .................... | .................... | .................... | .................... | .................... | |
| D | 6536 hCoV-19/Scotland/EDB4962/2020\|EPI_ISL_473810\|2020- | .................... | .................... | .................... | .................... | .................... | .................... | |
| D | 6537 hCoV-19/Scotland/EDB4965/2020\|EPI_ISL_473811\|2020- | .................... | .................... | .................... | .................... | .................... | .................... | |
| D | 6538 hCoV-19/Scotland/EDB4966/2020\|EPI_ISL_473812\|2020- | .................... | .................... | .................... | .................... | .................... | .................... | |
| D | 6539 hCoV-19/Scotland/EDB5157/2020\|EPI_ISL_473813\|2020- | .................... | .................... | .................... | .................... | .................... | .................... | |
| D | 6540 hCoV-19/Scotland/EDB5165/2020\|EPI_ISL_473814\|2020- | .................... | .A.................. | .................... | .................... | .................... | .................... | |
| D | 6541 hCoV-19/Scotland/EDB5196/2020\|EPI_ISL_473815\|2020- | .................... | .................... | .................... | .................... | .................... | .................... | |
| D | 6542 hCoV-19/Scotland/EDB5435/2020\|EPI_ISL_473817\|2020- | .................... | ...................A | .................... | .................... | .................... | .................... | 1 |
| D | 6543 hCoV-19/Scotland/EDB5436/2020\|EPI_ISL_473818\|2020- | .................... | ...................A | .................... | .................... | .................... | .................... | 1 |
| D | 6544 hCoV-19/Scotland/EDB5437/2020\|EPI_ISL_473819\|2020- | .................... | ...A................ | .................... | .................... | .................... | .................... | |
| D | 6545 hCoV-19/Scotland/EDB5438/2020\|EPI_ISL_473820\|2020- | .................... | ...................A | .................... | .................... | .................... | .................... | 1 |
| D | 6546 hCoV-19/Scotland/EDB5441/2020\|EPI_ISL_473821\|2020- | .................... | .................... | .................... | .................... | .................... | .................... | |
| D | 6547 hCoV-19/Scotland/EDB5442/2020\|EPI_ISL_473822\|2020- | .................... | ...................A | .................... | .................... | .................... | .................... | 1 |
| D | 6548 hCoV-19/Scotland/EDB5443/2020\|EPI_ISL_473823\|2020- | .................... | ...................A | .................... | .................... | .................... | .................... | 1 |
| D | 6549 hCoV-19/Scotland/EDB5444/2020\|EPI_ISL_473824\|2020- | .................... | ...................A | .................... | .................... | .................... | .................... | 1 |
| D | 6550 hCoV-19/Scotland/EDB5661/2020\|EPI_ISL_473825\|2020- | .................... | .................... | .................... | .................... | .................... | .................... | |
| D | 6551 hCoV-19/Scotland/EDB5667/2020\|EPI_ISL_473826\|2020- | .................... | ...................A | .................... | .................... | .................... | .................... | 1 |
| D | 6552 hCoV-19/Scotland/EDB5668/2020\|EPI_ISL_473827\|2020- | .................... | .................... | .................... | .................... | .................... | .................... | |
| D | 6553 hCoV-19/Scotland/EDB5669/2020\|EPI_ISL_473828\|2020- | .................... | .................... | .................... | .................... | .................... | .................... | |
| D | 6554 hCoV-19/Scotland/EDB5671/2020\|EPI_ISL_473829\|2020- | .................... | .................... | .................... | .................... | .................... | .................... | |
| D | 6555 hCoV-19/Scotland/EDB5672/2020\|EPI_ISL_473830\|2020- | .................... | ...................A | .................... | .................... | .................... | .................... | 1 |
| D | 6556 hCoV-19/Scotland/EDB5673/2020\|EPI_ISL_473831\|2020- | .................... | ...................A | .................... | .................... | .................... | .................... | 1 |
| D | 6557 hCoV-19/Scotland/EDB5676/2020\|EPI_ISL_473832\|2020- | .................... | .A.................. | .................... | .................... | .................... | .................... | |
| D | 6558 hCoV-19/Scotland/EDB5678/2020\|EPI_ISL_473833\|2020- | .................... | .................... | .................... | .................... | .................... | .................... | |
| D | 6559 hCoV-19/Scotland/EDB5680/2020\|EPI_ISL_473834\|2020- | .................... | .A.................. | .................... | .................... | .................... | .................... | |
| D | 6560 hCoV-19/Scotland/EDB5683/2020\|EPI_ISL_473836\|2020- | .................... | .................... | .................... | .................... | .................... | .................... | |
| D | 6561 hCoV-19/Scotland/EDB5710/2020\|EPI_ISL_473838\|2020- | .................... | ...................A | .................... | .................... | .................... | .................... | 1 |

| | | | | | | | | |
|---|---|---|---|---|---|---|---|---|
| D | 6562 hCoV-19/Scotland/EDB5711/2020\|EPI_ISL_473839\|2020- | .................... | .................... | .................... | .................... | .................... | .................... | |
| D | 6563 hCoV-19/Scotland/EDB5712/2020\|EPI_ISL_473840\|2020- | .................... | ...................A | .................... | .................... | .................... | .................... | 1 |
| D | 6564 hCoV-19/Scotland/EDB5713/2020\|EPI_ISL_473841\|2020- | .................... | .................... | .................... | .................... | .................... | .................... | |
| D | 6565 hCoV-19/Scotland/EDB5723/2020\|EPI_ISL_473844\|2020- | .................... | .................... | .................... | .................... | .................... | .................... | |
| D | 6566 hCoV-19/Scotland/EDB5729/2020\|EPI_ISL_473846\|2020- | .................... | ...................A | .................... | .................... | .................... | .................... | 1 |
| D | 6567 hCoV-19/Scotland/EDB5730/2020\|EPI_ISL_473847\|2020- | .................... | .................... | .................... | .................... | .................... | .................... | |
| D | 6568 hCoV-19/Scotland/EDB5731/2020\|EPI_ISL_473848\|2020- | .................... | .................... | .................... | .................... | .................... | .................... | |
| D | 6569 hCoV-19/Scotland/EDB5732/2020\|EPI_ISL_473849\|2020- | .................... | .................... | .................... | .................... | .................... | .................... | |
| D | 6570 hCoV-19/Scotland/EDB5733/2020\|EPI_ISL_473850\|2020- | .................... | .................... | .................... | .................... | .................... | .................... | |
| D | 6571 hCoV-19/Scotland/EDB5735/2020\|EPI_ISL_473851\|2020- | .................... | .................... | .................... | .................... | .................... | .................... | |
| D | 6572 hCoV-19/Scotland/EDB5745/2020\|EPI_ISL_473852\|2020- | .................... | .................... | .................... | .................... | .................... | .................... | |
| D | 6573 hCoV-19/Scotland/EDB5747/2020\|EPI_ISL_473854\|2020- | .................... | .................... | .................... | .................... | .................... | .................... | |
| D | 6574 hCoV-19/Scotland/EDB5780/2020\|EPI_ISL_473855\|2020- | .................... | ...................A | .................... | .................... | .................... | .................... | 1 |
| D | 6575 hCoV-19/Scotland/EDB5792/2020\|EPI_ISL_473856\|2020- | .................... | ...................A | .................... | .................... | .................... | .................... | 1 |
| D | 6576 hCoV-19/Scotland/EDB5793/2020\|EPI_ISL_473857\|2020- | .................... | ...................A | .................... | .................... | .................... | .................... | 1 |
| D | 6577 hCoV-19/Scotland/EDB5797/2020\|EPI_ISL_473858\|2020- | .................... | ...................A | .................... | .................... | .................... | .................... | 1 |
| D | 6578 hCoV-19/Scotland/EDB5799/2020\|EPI_ISL_473859\|2020- | .................... | ...................A | .................... | .................... | .................... | .................... | 1 |
| D | 6579 hCoV-19/Scotland/EDB5806/2020\|EPI_ISL_473860\|2020- | .................... | ...................A | .................... | .................... | .................... | .................... | 1 |
| D | 6581 hCoV-19/Scotland/EDB5842/2020\|EPI_ISL_473862\|2020- | .................... | ...................A | .................... | .................... | .................... | .................... | 1 |
| D | 6582 hCoV-19/Scotland/EDB5852/2020\|EPI_ISL_473863\|2020- | .................... | ...................A | .................... | .................... | .................... | .................... | 1 |
| D | 6583 hCoV-19/Scotland/EDB5853/2020\|EPI_ISL_473864\|2020- | .................... | ...................A | .................... | .................... | .................... | .................... | 1 |
| D | 6584 hCoV-19/Scotland/EDB5875/2020\|EPI_ISL_473865\|2020- | .................... | .................... | .................... | .................... | .................... | .................... | |
| D | 6586 hCoV-19/Scotland/EDB5892/2020\|EPI_ISL_473867\|2020- | .................... | ...................A | .................... | .................... | .................... | .................... | 1 |
| D | 6587 hCoV-19/Scotland/EDB5893/2020\|EPI_ISL_473868\|2020- | .................... | ...................A | .................... | .................... | .................... | .................... | 1 |
| D | 6588 hCoV-19/Scotland/EDB5894/2020\|EPI_ISL_473869\|2020- | .................... | ...................A | .................... | .................... | .................... | .................... | 1 |
| D | 6589 hCoV-19/Scotland/EDB5900/2020\|EPI_ISL_473870\|2020- | .................... | ...................A | .................... | .................... | .................... | .................... | 1 |
| D | 6590 hCoV-19/Scotland/EDB5941/2020\|EPI_ISL_473871\|2020- | .................... | ...................A | .................... | .................... | .................... | .................... | 1 |
| D | 6592 hCoV-19/Scotland/EDB6127/2020\|EPI_ISL_473873\|2020- | .................... | ...................A | .................... | .................... | .................... | .................... | 1 |
| D | 6593 hCoV-19/Scotland/EDB6128/2020\|EPI_ISL_473874\|2020- | .................... | ...................A | .................... | .................... | .................... | .................... | 1 |
| D | 6594 hCoV-19/Scotland/EDB6129/2020\|EPI_ISL_473875\|2020- | .................... | .A.................. | .................... | .................... | .................... | .................... | |
| D | 6595 hCoV-19/Scotland/EDB6130/2020\|EPI_ISL_473876\|2020- | .................... | .A.................. | .................... | .................... | .................... | .................... | |
| D | 6596 hCoV-19/Scotland/EDB6131/2020\|EPI_ISL_473877\|2020- | .................... | ...................A | .................... | .................... | .................... | .................... | 1 |
| D | 6597 hCoV-19/Scotland/EDB6133/2020\|EPI_ISL_473878\|2020- | .................... | .................... | .................... | .................... | .................... | .................... | |
| D | 6598 hCoV-19/Scotland/EDB6136/2020\|EPI_ISL_473880\|2020- | .................... | .................... | .................... | .................... | .................... | .................... | |
| D | 6599 hCoV-19/Scotland/EDB6138/2020\|EPI_ISL_473881\|2020- | .................... | .................... | .................... | .................... | .................... | .................... | |
| D | 6600 hCoV-19/USA/MI-MDHHS-SC20901/2020\|EPI_ISL_476038\|2 | .................... | .................... | .................... | .................... | .................... | .................... | |
| D | 6601 hCoV-19/USA/MI-MDHHS-SC20902/2020\|EPI_ISL_476039\|2 | .................... | .................... | .................... | .................... | .................... | .................... | |
| D | 6602 hCoV-19/USA/MI-MDHHS-SC20903/2020\|EPI_ISL_476040\|2 | .................... | .................... | .................... | .................... | .................... | .................... | |
| D | 6603 hCoV-19/USA/MI-MDHHS-SC20905/2020\|EPI_ISL_476041\|2 | .................... | .................... | .................... | .................... | .................... | .................... | |
| D | 6604 hCoV-19/USA/MI-MDHHS-SC20906/2020\|EPI_ISL_476042\|2 | .................... | .................... | .................... | .................... | .................... | .................... | |
| D | 6605 hCoV-19/USA/MI-MDHHS-SC20907/2020\|EPI_ISL_476043\|2 | .................... | .................... | .................... | .................... | .................... | .................... | |
| D | 6606 hCoV-19/USA/MI-MDHHS-SC20908/2020\|EPI_ISL_476044\|2 | .................... | .................... | .................... | .................... | .................... | .................... | |
| D | 6607 hCoV-19/USA/MI-MDHHS-SC20909/2020\|EPI_ISL_476045\|2 | .................... | .................... | .................... | .................... | .................... | .................... | |
| D | 6608 hCoV-19/USA/MI-MDHHS-SC20910/2020\|EPI_ISL_476046\|2 | .................... | .................... | .................... | .................... | .................... | .................... | |
| D | 6609 hCoV-19/USA/MI-MDHHS-SC20912/2020\|EPI_ISL_476048\|2 | .................... | .................... | .................... | .................... | .................... | .................... | |
| D | 6610 hCoV-19/USA/MI-MDHHS-SC20913/2020\|EPI_ISL_476049\|2 | .................... | .................... | .................... | .................... | .................... | .................... | |
| D | 6611 hCoV-19/USA/MI-MDHHS-SC20914/2020\|EPI_ISL_476050\|2 | .................... | .................... | .................... | .................... | .................... | .................... | |
| D | 6612 hCoV-19/USA/MI-MDHHS-SC20915/2020\|EPI_ISL_476051\|2 | .................... | .................... | .................... | .................... | .................... | .................... | |
| D | 6613 hCoV-19/USA/MI-MDHHS-SC20917/2020\|EPI_ISL_476053\|2 | .................... | .................... | .................... | .................... | .................... | .................... | |
| D | 6614 hCoV-19/USA/MI-MDHHS-SC20918/2020\|EPI_ISL_476054\|2 | .................... | .................... | .................... | .................... | .................... | .................... | |
| D | 6615 hCoV-19/USA/MI-MDHHS-SC20919/2020\|EPI_ISL_476055\|2 | .................... | .................... | .................... | .................... | .................... | .................... | |
| D | 6616 hCoV-19/USA/MI-MDHHS-SC20920/2020\|EPI_ISL_476056\|2 | .................... | .................... | .................... | .................... | .................... | .................... | |
| D | 6617 hCoV-19/USA/MI-MDHHS-SC20922/2020\|EPI_ISL_476058\|2 | .................... | .................... | .................... | .................... | .................... | .................... | |
| D | 6618 hCoV-19/USA/MI-MDHHS-SC20923/2020\|EPI_ISL_476059\|2 | .................... | .................... | .................... | .................... | .................... | .................... | |
| D | 6619 hCoV-19/USA/MI-MDHHS-SC20924/2020\|EPI_ISL_476060\|2 | .................... | .................... | .................... | .................... | .................... | .................... | |
| D | 6620 hCoV-19/USA/MI-MDHHS-SC20926/2020\|EPI_ISL_476062\|2 | .................... | .................... | .................... | .................... | .................... | .................... | |
| D | 6621 hCoV-19/USA/MI-MDHHS-SC20927/2020\|EPI_ISL_476063\|2 | .................... | .................... | .................... | .................... | .................... | .................... | |
| D | 6622 hCoV-19/USA/MI-MDHHS-SC20928/2020\|EPI_ISL_476064\|2 | .................... | .................... | .................... | .................... | .................... | .................... | |
| D | 6623 hCoV-19/Czech Republic/NRL_4033/2020\|EPI_ISL_47606 | .................... | .................... | .................... | .................... | .................... | .................... | |
| D | 6624 hCoV-19/Hungary/UD-10261/2020\|EPI_ISL_476068\|2020- | .................... | .................... | .................... | .................... | .................... | .................... | |
| D | 6625 hCoV-19/Hungary/UD-5394/2020\|EPI_ISL_476069\|2020-0 | .................... | .................... | .................... | .................... | .................... | .................... | |
| D | 6626 hCoV-19/Hungary/UD-6278/2020\|EPI_ISL_476070\|2020-0 | .................... | .................... | .................... | .................... | .................... | .................... | |
| D | 6627 hCoV-19/Hungary/UD-6858/2020\|EPI_ISL_476071\|2020-0 | .................... | .................... | .................... | .................... | .................... | .................... | |
| D | 6628 hCoV-19/Hungary/UD-7039/2020\|EPI_ISL_476072\|2020-0 | .................... | .................... | .................... | .................... | .................... | .................... | |
| D | 6629 hCoV-19/Hungary/UD-7040/2020\|EPI_ISL_476073\|2020-0 | .................... | .................... | .................... | .................... | .................... | .................... | |
| D | 6630 hCoV-19/Hungary/UD-7373/2020\|EPI_ISL_476074\|2020-0 | .................... | .................... | .................... | .................... | .................... | .................... | |
| D | 6631 hCoV-19/Hungary/UD-7417/2020\|EPI_ISL_476075\|2020-0 | .................... | .................... | .................... | .................... | .................... | .................... | |
| D | 6632 hCoV-19/Hungary/UD-7729/2020\|EPI_ISL_476076\|2020-0 | .................... | .................... | .................... | .................... | .................... | .................... | |
| D | 6633 hCoV-19/Hungary/UD-8157/2020\|EPI_ISL_476077\|2020-0 | .................... | .................... | .................... | .................... | .................... | .................... | |
| D | 6634 hCoV-19/Hungary/US-636w/2020\|EPI_ISL_476078\|2020-0 | .................... | .................... | .................... | .................... | .................... | .................... | |
| D | 6635 hCoV-19/Switzerland/BL-ETHZ-140000/2020\|EPI_ISL_47 | .................... | .................... | .................... | .................... | .................... | .................... | |
| D | 6636 hCoV-19/Switzerland/ZH-ETHZ-140001/2020\|EPI_ISL_47 | .................... | .................... | .................... | .................... | ......S............. | .................... | |
| D | 6637 hCoV-19/Switzerland/ZH-ETHZ-140002/2020\|EPI_ISL_47 | .................... | .................... | .................... | .................... | .................... | .................... | |
| D | 6638 hCoV-19/Switzerland/BE-ETHZ-140003/2020\|EPI_ISL_47 | .................... | .................... | .................... | .................... | ......S............. | .................... | |
| D | 6639 hCoV-19/Switzerland/BE-ETHZ-140004/2020\|EPI_ISL_47 | .................... | .A.................. | .................... | .................... | ......S............. | .................... | |
| D | 6640 hCoV-19/Switzerland/SO-ETHZ-140008/2020\|EPI_ISL_47 | .................... | .................... | .................... | .................... | ......M............. | .................... | |
| D | 6641 hCoV-19/Switzerland/SO-ETHZ-140009/2020\|EPI_ISL_47 | .................... | .................... | .................... | .................... | .................... | .................... | |
| D | 6642 hCoV-19/Switzerland/LU-ETHZ-140011/2020\|EPI_ISL_47 | .................... | .................... | .................... | .................... | .................... | .................... | |

| | | | | | | | |
|---|---|---|---|---|---|---|---|
| D | 6643 hCoV-19/Switzerland/BE-ETHZ-140017/2020\|EPI_ISL_47 | ···················· | ···················· | ···················· | ···················· | ·······S············ | ······R············· |
| D | 6644 hCoV-19/Switzerland/SG-ETHZ-140018/2020\|EPI_ISL_47 | ···················· | ···················· | ···················· | ···················· | ···················· | ···················· |
| D | 6645 hCoV-19/Switzerland/TI-ETHZ-140019/2020\|EPI_ISL_47 | ···················· | ···················· | ···················· | ···················· | ···················· | ···················· |
| D | 6646 hCoV-19/Switzerland/BL-ETHZ-140020/2020\|EPI_ISL_47 | ···················· | ···················· | ···················· | ···················· | ·······S············ | ···················· |
| D | 6647 hCoV-19/Switzerland/VD-ETHZ-140025/2020\|EPI_ISL_47 | ···················· | ···················· | ···················· | ···················· | ···················· | ···················· |
| D | 6648 hCoV-19/Switzerland/ZH-ETHZ-140028/2020\|EPI_ISL_47 | ···················· | ···················· | ···················· | ···················· | ···················· | ·········Y·········· |
| D | 6649 hCoV-19/Switzerland/BL-ETHZ-140032/2020\|EPI_ISL_47 | ···················· | ···················· | ···················· | ···················· | ·······S············ | ···················· |
| D | 6650 hCoV-19/Switzerland/BE-ETHZ-140033/2020\|EPI_ISL_47 | ···················· | ·A·················· | ···················· | ···················· | ·······S············ | ···················· |
| D | 6652 hCoV-19/Switzerland/BE-ETHZ-140043/2020\|EPI_ISL_47 | ···················· | ·A·················· | ···················· | ···················· | ···················· | ···················· |
| D | 6653 hCoV-19/Switzerland/TI-ETHZ-140047/2020\|EPI_ISL_47 | ···················· | ···················· | ···················· | ···················· | ···················· | ···················· |
| D | 6655 hCoV-19/England/OXON-B0694/2020\|EPI_ISL_478898\|202 | ···················· | ···················· | ···················· | ···················· | ···················· | ···················· |
| D | 6656 hCoV-19/England/OXON-B06A3/2020\|EPI_ISL_478899\|202 | ···················· | ···················· | ···················· | ···················· | ···················· | ···················· |
| D | 6657 hCoV-19/England/20099000504/2020\|EPI_ISL_464200\|20 | ···················· | ···················· | ···················· | ···················· | ···················· | ···················· |
| D | 6658 hCoV-19/England/20099000804/2020\|EPI_ISL_464201\|20 | ···················· | ···················· | ···················· | ···················· | ···················· | ····N··········C···· |
| D | 6659 hCoV-19/England/20099015104/2020\|EPI_ISL_464202\|20 | ···················· | ···················· | ···················· | ···················· | ···················· | ···················· |
| D | 6660 hCoV-19/England/20099044904/2020\|EPI_ISL_464204\|20 | ···················· | ···················· | ···················· | ···················· | ···················· | ···················· |
| D | 6661 hCoV-19/England/20099048504/2020\|EPI_ISL_464206\|20 | ···················· | ···················· | ···················· | ···················· | ···················· | ···················· |
| D | 6664 hCoV-19/England/20099066204/2020\|EPI_ISL_464209\|20 | ···················· | ···················· | ···················· | ···················· | ···················· | ···················· |
| D | 6665 hCoV-19/England/20099068104/2020\|EPI_ISL_464210\|20 | ···················· | ···················· | ···················· | ···················· | ···················· | ···················· |
| D | 6666 hCoV-19/England/20099068604/2020\|EPI_ISL_464212\|20 | ···················· | ···················· | ···················· | ···················· | ···················· | ···················· |
| D | 6669 hCoV-19/England/20102000704/2020\|EPI_ISL_464262\|20 | ···················· | ···················· | ···················· | ···················· | ···················· | ···················· |
| D | 6671 hCoV-19/England/20102001704/2020\|EPI_ISL_464270\|20 | ···················· | ···················· | ···················· | ···················· | ···················· | ···················· |
| D | 6672 hCoV-19/England/20102003804/2020\|EPI_ISL_464275\|20 | ···················· | ···················· | ···················· | ···················· | ···················· | ···················· |
| D | 6673 hCoV-19/England/20102004104/2020\|EPI_ISL_464278\|20 | ···················· | ···················· | ···················· | ···················· | ···················· | ···················· |
| D | 6674 hCoV-19/England/20102005304/2020\|EPI_ISL_464283\|20 | ···················· | ···················· | ···················· | ···················· | ···················· | ···················· |
| D | 6676 hCoV-19/England/20102133304/2020\|EPI_ISL_464291\|20 | ···················· | ···················· | ···················· | ···················· | ···················· | ···················· |
| D | 6678 hCoV-19/England/201021554/2020\|EPI_ISL_464293\|2020 | ···················· | ···················· | ···················· | ···················· | ···················· | ···················· |
| D | 6679 hCoV-19/England/201021555/2020\|EPI_ISL_464294\|2020 | ···················· | ···················· | ···················· | ···················· | ···················· | ···················· |
| D | 6680 hCoV-19/England/201021557/2020\|EPI_ISL_464295\|2020 | ···················· | ···················· | ···················· | ············R······· | ···················· | ···················· |
| D | 6681 hCoV-19/England/201021559/2020\|EPI_ISL_464296\|2020 | ···················· | ···················· | ···················· | ···················· | ···················· | ···················· |
| D | 6682 hCoV-19/England/201021560/2020\|EPI_ISL_464297\|2020 | ···················· | ···················· | ···················· | ···················· | ···················· | ····N··············· |
| D | 6687 hCoV-19/England/20188035504/2020\|EPI_ISL_465505\|20 | ···················· | ···················· | ···················· | ···················· | ···················· | ···················· |
| D | 6689 hCoV-19/England/20188051804/2020\|EPI_ISL_465508\|20 | ···················· | ···················· | ···················· | ···················· | ···················· | ···················· |
| D | 6690 hCoV-19/England/20188064104/2020\|EPI_ISL_465510\|20 | ···················· | ···················· | ···················· | ···················· | ···················· | ···················· |
| D | 6691 hCoV-19/England/20188072401/2020\|EPI_ISL_465513\|20 | ···················· | ···················· | ···················· | ···················· | ···················· | ···················· |
| D | 6692 hCoV-19/England/20188072804/2020\|EPI_ISL_465514\|20 | ···················· | ···················· | ···················· | ···················· | ···················· | ···················· |
| D | 6693 hCoV-19/England/20188142804/2020\|EPI_ISL_465517\|20 | ···················· | ···················· | ···················· | ···················· | ···················· | ···················· |
| D | 6698 hCoV-19/England/20189019804/2020\|EPI_ISL_465525\|20 | ···················· | ···················· | ···················· | ···················· | ···················· | ···················· |
| D | 6699 hCoV-19/England/20189026604/2020\|EPI_ISL_465526\|20 | ···················· | ···················· | ···················· | ···················· | ···················· | ···················· |
| D | 6702 hCoV-19/England/20189046701/2020\|EPI_ISL_465531\|20 | ···················· | ···················· | ···················· | ···················· | ···················· | ···················· |
| D | 6704 hCoV-19/England/20189050701/2020\|EPI_ISL_465533\|20 | ···················· | ···················· | ···················· | ···················· | ···················· | ···················· |
| D | 6705 hCoV-19/England/20189051801/2020\|EPI_ISL_465534\|20 | ···················· | ···················· | ···················· | ···················· | ···················· | ···················· |
| D | 6707 hCoV-19/England/20189054401/2020\|EPI_ISL_465537\|20 | ···················· | ···················· | ···················· | ···················· | ···················· | ···················· |
| D | 6708 hCoV-19/England/20189080701/2020\|EPI_ISL_465539\|20 | ···················· | ···················· | ···················· | ···················· | ···················· | ···················· |
| D | 6710 hCoV-19/England/20189126801/2020\|EPI_ISL_465546\|20 | ···················· | ···················· | ···················· | ···················· | ···················· | ···················· |
| D | 6712 hCoV-19/England/20190056901/2020\|EPI_ISL_465589\|20 | ···················· | ···················· | ···················· | ···················· | ···················· | ···················· |
| D | 6713 hCoV-19/England/20190062001/2020\|EPI_ISL_465590\|20 | ···················· | ···················· | ···················· | ···················· | ···················· | ···················· |
| D | 6715 hCoV-19/England/20190141101/2020\|EPI_ISL_465592\|20 | ···················· | ···················· | ···················· | ···················· | ···················· | ···················· |
| D | 6718 hCoV-19/England/20192004301/2020\|EPI_ISL_465595\|20 | ···················· | ···················· | ···················· | ···················· | ···················· | ···················· |
| D | 6720 hCoV-19/England/20192018201/2020\|EPI_ISL_465597\|20 | ···················· | ···················· | ···················· | ···················· | ···················· | ···················· |
| D | 6721 hCoV-19/England/20192020301/2020\|EPI_ISL_465598\|20 | ···················· | ···················· | ···················· | ···················· | ···················· | ···················· |
| D | 6722 hCoV-19/Canada/BC_06378958/2020\|EPI_ISL_466800\|202 | ···················· | ···················· | ···················· | ···················· | ···················· | ···················· |
| D | 6723 hCoV-19/Canada/BC_06764319/2020\|EPI_ISL_466802\|202 | ···················· | ···················· | ···················· | ···················· | ···················· | ···················· |
| D | 6724 hCoV-19/Canada/BC_06768018/2020\|EPI_ISL_466803\|202 | ···················· | ···················· | ···················· | ···················· | ···················· | ···················· |
| D | 6725 hCoV-19/Canada/BC_06803213/2020\|EPI_ISL_466806\|202 | ···················· | ···················· | ···················· | ···················· | ···················· | ···················· |
| D | 6726 hCoV-19/Canada/BC_06852453/2020\|EPI_ISL_466807\|202 | ···················· | ···················· | ···················· | ···················· | ···················· | ···················· |
| D | 6727 hCoV-19/Canada/BC_06944842/2020\|EPI_ISL_466808\|202 | ···················· | ···················· | ···················· | ···················· | ···················· | ···················· |
| D | 6728 hCoV-19/Canada/BC_06946792/2020\|EPI_ISL_466809\|202 | ···················· | ···················· | ···················· | ···················· | ···················· | ···················· |
| D | 6729 hCoV-19/Canada/BC_06955362/2020\|EPI_ISL_466810\|202 | ···················· | ···················· | ···················· | ···················· | ···················· | ···················· |
| D | 6730 hCoV-19/Canada/BC_06975953/2020\|EPI_ISL_466811\|202 | ···················· | ·A·················· | ···················· | ···················· | ···················· | ···················· |
| D | 6731 hCoV-19/Canada/BC_06998971/2020\|EPI_ISL_466812\|202 | ···················· | ···················· | ···················· | ···················· | ···················· | ···················· |
| D | 6732 hCoV-19/Canada/BC_07229190/2020\|EPI_ISL_466814\|202 | ···················· | ···················· | ···················· | ···················· | ···················· | ···················· |
| D | 6733 hCoV-19/Canada/BC_07312838/2020\|EPI_ISL_466815\|202 | ···················· | ···················· | ···················· | ···················· | ···················· | ···················· |
| D | 6734 hCoV-19/Canada/BC_07360020/2020\|EPI_ISL_466816\|202 | ···················· | ···················· | ···················· | ···················· | ···················· | ···················· |
| D | 6735 hCoV-19/Canada/BC_07720149/2020\|EPI_ISL_466819\|202 | ···················· | ···················· | ···················· | ···················· | ···················· | ···················· |
| D | 6736 hCoV-19/Canada/BC_07788742/2020\|EPI_ISL_466820\|202 | ···················· | ···················· | ···················· | ···················· | ···················· | ···················· |
| D | 6737 hCoV-19/Canada/BC_07812071/2020\|EPI_ISL_466821\|202 | ···················· | ···················· | ···················· | ···················· | ···················· | ···················· |
| D | 6738 hCoV-19/Canada/BC_07833956/2020\|EPI_ISL_466822\|202 | ···················· | ···················· | ···················· | ···················· | ···················· | ···················· |
| D | 6739 hCoV-19/Canada/BC_07884511/2020\|EPI_ISL_466824\|202 | ···················· | ···················· | ···················· | ···················· | ···················· | ···················· |
| D | 6740 hCoV-19/Canada/BC_07986535/2020\|EPI_ISL_466826\|202 | ···················· | ···················· | ···················· | ···················· | ···················· | ···················· |
| D | 6741 hCoV-19/Canada/BC_08323846/2020\|EPI_ISL_466829\|202 | ···················· | ···················· | ···················· | ···················· | ···················· | ···················· |
| D | 6742 hCoV-19/Canada/BC_08347845/2020\|EPI_ISL_466831\|202 | ···················· | ···················· | ···················· | ···················· | ···················· | ···················· |
| D | 6743 hCoV-19/Canada/BC_08419582/2020\|EPI_ISL_466832\|202 | ···················· | ···················· | ···················· | ···················· | ···················· | ···················· |
| D | 6744 hCoV-19/Canada/BC_08847088/2020\|EPI_ISL_466834\|202 | ···················· | ···················· | ···················· | ···················· | ···················· | ···················· |
| D | 6745 hCoV-19/Canada/BC_08940467/2020\|EPI_ISL_466838\|202 | ···················· | ···················· | ···················· | ···················· | ···················· | ···················· |
| D | 6746 hCoV-19/India/TG-NGC-CDFD-01/2020\|EPI_ISL_466839\|2 | ···················· | ···················· | ···················· | ···················· | ···················· | ···················· |
| D | 6747 hCoV-19/India/TG-NGC-CDFD-03/2020\|EPI_ISL_466841\|2 | ···················· | ···················· | ···················· | ···················· | ···················· | ···················· |
| D | 6748 hCoV-19/India/TG-NGC-CDFD-12/2020\|EPI_ISL_466850\|2 | ···················· | ···················· | ···················· | ···················· | ···················· | ···················· |

| | | | | | | | |
|---|---|---|---|---|---|---|---|
| D | 6749 hCoV-19/India/TG-NGC-CDFD-15/2020\|EPI_ISL_466853\|2 | .................... | .................... | .................... | .................... | .................... | .................... |
| D | 6750 hCoV-19/India/TG-NGC-CDFD-17/2020\|EPI_ISL_466855\|2 | .................... | .................... | .................... | .................... | .................... | .................... |
| D | 6751 hCoV-19/India/TG-NGC-CDFD-19/2020\|EPI_ISL_466857\|2 | .................... | .................... | .................... | .................... | .................... | .................... |
| D | 6752 hCoV-19/India/TG-NGC-CDFD-22/2020\|EPI_ISL_466860\|2 | .................... | .................... | .................... | .................... | .................... | .................... |
| D | 6753 hCoV-19/India/TG-NGC-CDFD-25/2020\|EPI_ISL_466863\|2 | .................... | .................... | .................... | .................... | .................... | .................... |
| D | 6754 hCoV-19/India/TG-NGC-CDFD-26/2020\|EPI_ISL_466864\|2 | .................... | .................... | .................... | .................... | .................... | .................... |
| D | 6755 hCoV-19/India/TG-NGC-CDFD-28/2020\|EPI_ISL_466866\|2 | .................... | .................... | .................... | .................... | .................... | .................... |
| D | 6756 hCoV-19/India/TG-NGC-CDFD-29/2020\|EPI_ISL_466867\|2 | .................... | .................... | .................... | .................... | .................... | .................... |
| D | 6757 hCoV-19/India/TG-NGC-CDFD-32/2020\|EPI_ISL_466870\|2 | .................... | .................... | .................... | .................... | .................... | .................... |
| D | 6758 hCoV-19/India/TG-NGC-CDFD-33/2020\|EPI_ISL_466871\|2 | .................... | .................... | .................... | .................... | .................... | .................... |
| D | 6759 hCoV-19/India/TG-NGC-CDFD-34/2020\|EPI_ISL_466872\|2 | .................... | .................... | .................... | .................... | .................... | .................... |
| D | 6760 hCoV-19/England/MZ001/2020\|EPI_ISL_466873\|2020-04 | .................... | .................... | .................... | .................... | .................... | .................... |
| D | 6761 hCoV-19/Germany/BY-MVP-0185/2020\|EPI_ISL_466878\|20 | .................... | .................... | .................... | .................... | .................... | .................... |
| D | 6762 hCoV-19/Germany/BY-MVP-0186/2020\|EPI_ISL_466879\|20 | .................... | .................... | .................... | .................... | .................... | .................... |
| D | 6763 hCoV-19/India/GJ-GBRC185a/2020\|EPI_ISL_469025\|2020 | .................... | .................... | .................... | .................... | .................... | .................... |
| D | 6764 hCoV-19/Sweden/20-51703/2020\|EPI_ISL_469070\|2020-0 | .................... | .................... | .................... | .................... | .................... | .................... |
| D | 6765 hCoV-19/Sweden/20-51758/2020\|EPI_ISL_469071\|2020-0 | .................... | .................... | .................... | .................... | .................... | .................... |
| D | 6766 hCoV-19/Sweden/20-51891/2020\|EPI_ISL_469072\|2020-0 | .................... | .................... | .................... | .................... | .................... | .................... |
| D | 6767 hCoV-19/Sweden/20-08130/2020\|EPI_ISL_469073\|2020-0 | .................... | .................... | .................... | .................... | .................... | .................... |
| D | 6768 hCoV-19/Sweden/20-08134/2020\|EPI_ISL_469074\|2020-0 | .................... | .................... | .................... | .................... | .................... | .................... |
| D | 6769 hCoV-19/Sweden/20-50106/2020\|EPI_ISL_469075\|2020-0 | .................... | .................... | .................... | .................... | .................... | .................... |
| D | 6770 hCoV-19/Sweden/20-50120/2020\|EPI_ISL_469076\|2020-0 | .................... | .................... | .................... | .................... | .................... | .................... |
| D | 6771 hCoV-19/Sweden/20-50216/2020\|EPI_ISL_469077\|2020-0 | .................... | .................... | .................... | .................... | .................... | .................... |
| D | 6772 hCoV-19/Sweden/20-50261/2020\|EPI_ISL_469078\|2020-0 | .................... | .................... | .................... | .................... | .................... | .................... |
| D | 6773 hCoV-19/Sweden/20-08089/2020\|EPI_ISL_469079\|2020-0 | .................... | .................... | .................... | .................... | .................... | .................... |
| D | 6774 hCoV-19/Singapore/327/2020\|EPI_ISL_469081\|2020-04- | .................... | .................... | .................... | .................... | .................... | .................... |
| D | 6775 hCoV-19/Singapore/328/2020\|EPI_ISL_469082\|2020-04- | .................... | .................... | .................... | .................... | .................... | .................... |
| D | 6776 hCoV-19/Singapore/329/2020\|EPI_ISL_469083\|2020-04- | .................... | .................... | .................... | .................... | .................... | .................... |
| D | 6777 hCoV-19/Singapore/330/2020\|EPI_ISL_469084\|2020-04- | .................... | .................... | .................... | .................... | .................... | .................... |
| D | 6778 hCoV-19/Singapore/331/2020\|EPI_ISL_469085\|2020-04- | .................... | .................... | .................... | .................... | .................... | .................... |
| D | 6779 hCoV-19/Singapore/332/2020\|EPI_ISL_469086\|2020-04- | .................... | .................... | .................... | .................... | .................... | .................... |
| D | 6780 hCoV-19/Singapore/333/2020\|EPI_ISL_469087\|2020-04- | .................... | .................... | .................... | .................... | .................... | .................... |
| D | 6781 hCoV-19/Singapore/334/2020\|EPI_ISL_469088\|2020-04- | .................... | .................... | .................... | .................... | .................... | .................... |
| D | 6782 hCoV-19/Singapore/335/2020\|EPI_ISL_469089\|2020-04- | .................... | .................... | .................... | .................... | .................... | .................... |
| D | 6783 hCoV-19/Singapore/336/2020\|EPI_ISL_469090\|2020-04- | .................... | .................... | .................... | .................... | .................... | .................... |
| D | 6784 hCoV-19/Singapore/337/2020\|EPI_ISL_469091\|2020-04- | .................... | .................... | .................... | .................... | .................... | .................... |
| D | 6785 hCoV-19/Singapore/338/2020\|EPI_ISL_469092\|2020-04- | .................... | .................... | .................... | .................... | .................... | .................... |
| D | 6786 hCoV-19/Singapore/339/2020\|EPI_ISL_469093\|2020-04- | .................... | .................... | .................... | .................... | .................... | .................... |
| D | 6787 hCoV-19/Singapore/340/2020\|EPI_ISL_469094\|2020-04- | .................... | .................... | .................... | .................... | .................... | .................... |
| D | 6788 hCoV-19/Singapore/341/2020\|EPI_ISL_469095\|2020-04- | .................... | .................... | .................... | .................... | .................... | .................... |
| D | 6789 hCoV-19/Singapore/342/2020\|EPI_ISL_469096\|2020-05- | .................... | .................... | .................... | .................... | .................... | .................... |
| D | 6790 hCoV-19/Singapore/344/2020\|EPI_ISL_469098\|2020-04- | .................... | .................... | .................... | .................... | .................... | .................... |
| D | 6791 hCoV-19/Singapore/345/2020\|EPI_ISL_469099\|2020-04- | .................... | .................... | .................... | .................... | .................... | .................... |
| D | 6792 hCoV-19/Northern Ireland/NIRE-FBB43/2020\|EPI_ISL_4 | .................... | .................... | .................... | .................... | .................... | .................... |
| D | 6793 hCoV-19/Northern Ireland/NIRE-FCB24/2020\|EPI_ISL_4 | .................... | .................... | .................... | .................... | .................... | .................... |
| D | 6794 hCoV-19/Northern Ireland/NIRE-FC201/2020\|EPI_ISL_4 | .................... | .................... | .................... | .................... | .................... | .................... |
| D | 6795 hCoV-19/Northern Ireland/NIRE-FCA90/2020\|EPI_ISL_4 | .................... | .................... | .................... | .................... | .................... | .................... |
| D | 6796 hCoV-19/Northern Ireland/NIRE-FB8B5/2020\|EPI_ISL_4 | .................... | .................... | .................... | .................... | .................... | .................... |
| D | 6797 hCoV-19/Northern Ireland/NIRE-FC104/2020\|EPI_ISL_4 | .................... | .................... | .................... | .................... | .................... | .................... |
| D | 6798 hCoV-19/Northern Ireland/NIRE-FC395/2020\|EPI_ISL_4 | .................... | .................... | .................... | .................... | .................... | .................... |
| D | 6799 hCoV-19/Northern Ireland/NIRE-FC18C/2020\|EPI_ISL_4 | .................... | .................... | .................... | .................... | .................... | .................... |
| D | 6800 hCoV-19/Northern Ireland/NIRE-FC5DB/2020\|EPI_ISL_4 | .................... | .................... | .................... | .................... | .................... | .................... |
| D | 6801 hCoV-19/Northern Ireland/NIRE-FBC8C/2020\|EPI_ISL_4 | .................... | .................... | .................... | .................... | .................... | .................... |
| D | 6802 hCoV-19/Northern Ireland/NIRE-FBCAA/2020\|EPI_ISL_4 | .................... | .................... | .................... | .................... | .................... | .................... |
| D | 6803 hCoV-19/Northern Ireland/NIRE-FBD2F/2020\|EPI_ISL_4 | .................... | .................... | .................... | .................... | .................... | .................... |
| D | 6804 hCoV-19/Northern Ireland/NIRE-FC76C/2020\|EPI_ISL_4 | .................... | .................... | .................... | .................... | .................... | .................... |
| D | 6805 hCoV-19/Northern Ireland/NIRE-FB85B/2020\|EPI_ISL_4 | .................... | .................... | .................... | .................... | .................... | .................... |
| D | 6806 hCoV-19/Northern Ireland/NIRE-FA704/2020\|EPI_ISL_4 | .................... | .................... | .................... | .................... | .................... | .................... |
| D | 6807 hCoV-19/Northern Ireland/NIRE-FBB16/2020\|EPI_ISL_4 | .................... | .................... | .................... | .................... | .................... | .................... |
| D | 6808 hCoV-19/Northern Ireland/NIRE-FBCB9/2020\|EPI_ISL_4 | .................... | .................... | .................... | .................... | .................... | .................... |
| D | 6809 hCoV-19/Northern Ireland/NIRE-FBA73/2020\|EPI_ISL_4 | .................... | .................... | .................... | .................... | .................... | .................... |
| D | 6810 hCoV-19/Northern Ireland/NIRE-FC447/2020\|EPI_ISL_4 | .................... | .................... | .................... | .................... | .................... | .................... |
| D | 6811 hCoV-19/Northern Ireland/NIRE-FBB70/2020\|EPI_ISL_4 | .................... | .................... | .................... | .................... | .................... | .................... |
| D | 6812 hCoV-19/Northern Ireland/NIRE-FAC41/2020\|EPI_ISL_4 | .................... | .................... | .................... | .................... | .................... | .................... |
| D | 6813 hCoV-19/England/CAMB-1AA5BD/2020\|EPI_ISL_470092\|20 | .................... | .................... | .................... | .................... | .................... | .................... |
| D | 6814 hCoV-19/England/CAMB-1ABE48/2020\|EPI_ISL_470096\|20 | .................... | .................... | .................... | .................... | .................... | .................... |
| D | 6815 hCoV-19/England/CAMB-1ABCA8/2020\|EPI_ISL_470098\|20 | .................... | .................... | .................... | .................... | .................... | .................... |
| D | 6816 hCoV-19/England/CAMB-1AB66E/2020\|EPI_ISL_470099\|20 | .................... | .................... | .................... | .................... | .................... | .................... |
| D | 6817 hCoV-19/DRC/3842/2020\|EPI_ISL_471396\|2020-04-25 | .................... | .................... | .................... | .................... | .................... | .................... |
| D | 6818 hCoV-19/DRC/3846/2020\|EPI_ISL_471397\|2020-04-25 | .................... | .................... | .................... | .................... | .................... | .................... |
| D | 6819 hCoV-19/DRC/3937/2020\|EPI_ISL_471398\|2020-04-25 | .................... | .................... | .................... | .................... | .................... | .................... |
| D | 6820 hCoV-19/Wales/PHWC-15D188/2020\|EPI_ISL_472676\|2020 | .................... | .................... | .................... | .................... | .................... | .................... |
| D | 6821 hCoV-19/Brazil/SP-L12-CD28/2020\|EPI_ISL_476221\|202 | .................... | .................... | .................... | .................... | .................... | .................... |
| D | 6822 hCoV-19/England/OXON-AFA9C/2020\|EPI_ISL_478801\|202 | .................... | .................... | .................... | .................... | .................... | .................... |
| D | 6823 hCoV-19/England/OXON-AFABA/2020\|EPI_ISL_478802\|202 | .................... | .................... | .................... | .................... | .................... | .................... |
| D | 6824 hCoV-19/England/OXON-AFAC9/2020\|EPI_ISL_478803\|202 | .................... | .................... | .................... | .................... | .................... | .................... |
| D | 6825 hCoV-19/England/OXON-AFAE7/2020\|EPI_ISL_478804\|202 | .................... | .................... | .................... | .................... | .................... | .................... |
| D | 6826 hCoV-19/England/OXON-AFB20/2020\|EPI_ISL_478805\|202 | .................... | .................... | .................... | .................... | .................... | .................... |

| | | | | | | | | | |
|---|---|---|---|---|---|---|---|---|---|
| D | 6827 | hCoV-19/England/OXON-AFB3F/2020\|EPI_ISL_478806\|202 | .................... | ...................A | .................... | .................... | .................... | .................... | 1 |
| D | 6828 | hCoV-19/England/OXON-AFB4E/2020\|EPI_ISL_478807\|202 | .................... | .................... | .................... | .................... | .................... | .................... | |
| D | 6829 | hCoV-19/England/OXON-AFB5D/2020\|EPI_ISL_478808\|202 | .................... | .................... | .................... | .................... | .................... | .................... | |
| D | 6830 | hCoV-19/England/OXON-AFB7B/2020\|EPI_ISL_478809\|202 | .................... | .................... | .................... | .................... | .................... | .................... | |
| D | 6831 | hCoV-19/England/OXON-AFBB7/2020\|EPI_ISL_478810\|202 | .................... | .................... | .................... | .................... | .................... | .................... | |
| D | 6832 | hCoV-19/England/OXON-AFBC6/2020\|EPI_ISL_478811\|202 | .................... | .................... | .................... | .................... | .................... | .................... | |
| D | 6833 | hCoV-19/England/OXON-AFBD5/2020\|EPI_ISL_478812\|202 | .................... | .................... | .................... | .................... | .................... | .................... | |
| D | 6834 | hCoV-19/England/OXON-AFC1E/2020\|EPI_ISL_478813\|202 | .................... | .................... | .................... | .................... | .................... | .................... | |
| D | 6835 | hCoV-19/England/OXON-AFC3C/2020\|EPI_ISL_478814\|202 | .................... | .................... | .................... | .................... | .................... | .................... | |
| D | 6836 | hCoV-19/England/OXON-AFC4B/2020\|EPI_ISL_478815\|202 | .................... | .................... | .................... | .................... | .................... | .................... | |
| D | 6837 | hCoV-19/England/OXON-AFC5A/2020\|EPI_ISL_478816\|202 | .................... | .................... | .................... | .................... | .................... | .................... | |
| D | 6838 | hCoV-19/England/OXON-AFC69/2020\|EPI_ISL_478817\|202 | .................... | .................... | .................... | .................... | .................... | .................... | |
| D | 6839 | hCoV-19/England/OXON-AFCA5/2020\|EPI_ISL_478818\|202 | .................... | .................... | .................... | .................... | .................... | .................... | |
| D | 6840 | hCoV-19/England/OXON-AFCB4/2020\|EPI_ISL_478819\|202 | .................... | .................... | .................... | .................... | .................... | .................... | |
| D | 6841 | hCoV-19/England/OXON-AFCD2/2020\|EPI_ISL_478820\|202 | .................... | .................... | .................... | .................... | .................... | .................... | |
| D | 6842 | hCoV-19/England/OXON-AFCE1/2020\|EPI_ISL_478821\|202 | .................... | .................... | .................... | .................... | .................... | .................... | |
| D | 6843 | hCoV-19/England/OXON-AFD0C/2020\|EPI_ISL_478822\|202 | .................... | .................... | .................... | .................... | .................... | .................... | |
| D | 6844 | hCoV-19/England/OXON-AFD1B/2020\|EPI_ISL_478823\|202 | .................... | .................... | .................... | .................... | .................... | .................... | |
| D | 6845 | hCoV-19/England/OXON-AFD57/2020\|EPI_ISL_478824\|202 | .................... | .................... | .................... | .................... | .................... | .................... | |
| D | 6846 | hCoV-19/England/OXON-AFD66/2020\|EPI_ISL_478825\|202 | .................... | .................... | .................... | .................... | .................... | .................... | |
| D | 6847 | hCoV-19/England/OXON-AFD75/2020\|EPI_ISL_478826\|202 | .................... | .................... | .................... | .................... | .................... | .................... | |
| D | 6848 | hCoV-19/England/OXON-AFDB1/2020\|EPI_ISL_478827\|202 | .................... | .................... | .................... | .................... | .................... | .................... | |
| D | 6849 | hCoV-19/England/OXON-AFDDF/2020\|EPI_ISL_478828\|202 | .................... | .................... | .................... | .................... | .................... | .................... | |
| D | 6850 | hCoV-19/England/OXON-AFE45/2020\|EPI_ISL_478829\|202 | .................... | .................... | .................... | .................... | .................... | .................... | |
| D | 6851 | hCoV-19/England/OXON-AFE54/2020\|EPI_ISL_478830\|202 | .................... | .................... | .................... | .................... | .................... | .................... | |
| D | 6852 | hCoV-19/England/OXON-AFE63/2020\|EPI_ISL_478831\|202 | .................... | .................... | .................... | .................... | .................... | .................... | |
| D | 6853 | hCoV-19/England/OXON-AFE90/2020\|EPI_ISL_478832\|202 | .................... | .................... | .................... | .................... | .................... | .................... | |
| D | 6854 | hCoV-19/England/OXON-AFEAF/2020\|EPI_ISL_478833\|202 | .................... | .................... | .................... | .................... | .................... | .................... | |
| D | 6855 | hCoV-19/England/OXON-AFEBE/2020\|EPI_ISL_478834\|202 | .................... | .................... | .................... | .................... | .................... | .................... | |
| D | 6856 | hCoV-19/England/OXON-AFEDC/2020\|EPI_ISL_478835\|202 | .................... | .................... | .................... | .................... | .................... | .................... | |
| D | 6857 | hCoV-19/England/OXON-AFF15/2020\|EPI_ISL_478836\|202 | .................... | .................... | .................... | .................... | .................... | .................... | |
| D | 6858 | hCoV-19/England/OXON-AFF42/2020\|EPI_ISL_478837\|202 | .................... | .................... | .................... | .................... | .................... | .................... | |
| D | 6859 | hCoV-19/England/OXON-AFF51/2020\|EPI_ISL_478838\|202 | .................... | .................... | .................... | .................... | .................... | .................... | |
| D | 6860 | hCoV-19/England/OXON-AFF7F/2020\|EPI_ISL_478839\|202 | .................... | .................... | .................... | .................... | .................... | .................... | |
| D | 6861 | hCoV-19/England/OXON-AFF8E/2020\|EPI_ISL_478840\|202 | .................... | .................... | .................... | .................... | .................... | .................... | |
| D | 6862 | hCoV-19/England/OXON-AFF9D/2020\|EPI_ISL_478841\|202 | .................... | .................... | .................... | .................... | .................... | .................... | |
| D | 6863 | hCoV-19/England/OXON-AFFCA/2020\|EPI_ISL_478842\|202 | .................... | .................... | .................... | .................... | .................... | .................... | |
| D | 6864 | hCoV-19/England/OXON-AFFD9/2020\|EPI_ISL_478843\|202 | .................... | .................... | .................... | .................... | .................... | .................... | |
| D | 6865 | hCoV-19/England/OXON-AFFF7/2020\|EPI_ISL_478844\|202 | .................... | .................... | .................... | .................... | .................... | .................... | |
| D | 6866 | hCoV-19/England/OXON-B001E/2020\|EPI_ISL_478845\|202 | .................... | .................... | .................... | .................... | .................... | .................... | |
| D | 6867 | hCoV-19/England/OXON-B002D/2020\|EPI_ISL_478846\|202 | .................... | .................... | .................... | .................... | .................... | .................... | |
| D | 6868 | hCoV-19/England/OXON-B003C/2020\|EPI_ISL_478847\|202 | .................... | .................... | .................... | .................... | .................... | .................... | |
| D | 6869 | hCoV-19/England/OXON-B005A/2020\|EPI_ISL_478848\|202 | .................... | .................... | .................... | .................... | .................... | .................... | |
| D | 6870 | hCoV-19/England/OXON-B0069/2020\|EPI_ISL_478849\|202 | .................... | .................... | .................... | .................... | .................... | .................... | |
| D | 6871 | hCoV-19/England/OXON-B0078/2020\|EPI_ISL_478850\|202 | .................... | .................... | .................... | .................... | .................... | .................... | |
| D | 6872 | hCoV-19/England/OXON-B0087/2020\|EPI_ISL_478851\|202 | .................... | .................... | .................... | .................... | .................... | .................... | |
| D | 6873 | hCoV-19/England/OXON-B0096/2020\|EPI_ISL_478852\|202 | .................... | .................... | .................... | .................... | .................... | .................... | |
| D | 6874 | hCoV-19/England/OXON-B00A5/2020\|EPI_ISL_478853\|202 | .................... | .................... | .................... | .................... | .................... | .................... | |
| D | 6875 | hCoV-19/England/OXON-B00D2/2020\|EPI_ISL_478854\|202 | .................... | .................... | .................... | .................... | .................... | .................... | |
| D | 6876 | hCoV-19/England/OXON-B00F0/2020\|EPI_ISL_478855\|202 | .................... | .................... | .................... | .................... | .................... | .................... | |
| D | 6877 | hCoV-19/England/OXON-B010C/2020\|EPI_ISL_478856\|202 | .................... | .................... | .................... | .................... | .................... | .................... | |
| D | 6878 | hCoV-19/England/OXON-B011B/2020\|EPI_ISL_478857\|202 | .................... | .................... | .................... | .................... | .................... | .................... | |
| D | 6879 | hCoV-19/England/OXON-B0175/2020\|EPI_ISL_478859\|202 | .................... | .................... | .................... | .................... | .................... | .................... | |
| D | 6880 | hCoV-19/England/OXON-B0184/2020\|EPI_ISL_478860\|202 | .................... | .................... | .................... | .................... | .................... | .................... | |
| D | 6881 | hCoV-19/England/OXON-B0193/2020\|EPI_ISL_478861\|202 | .................... | .................... | .................... | .................... | .................... | .................... | |
| D | 6882 | hCoV-19/England/OXON-B01A2/2020\|EPI_ISL_478862\|202 | .................... | .................... | .................... | .................... | .................... | .................... | |
| D | 6883 | hCoV-19/England/OXON-B01B1/2020\|EPI_ISL_478863\|202 | .................... | .................... | .................... | .................... | .................... | .................... | |
| D | 6884 | hCoV-19/England/OXON-B01DF/2020\|EPI_ISL_478864\|202 | .................... | .................... | .................... | .................... | .................... | .................... | |
| D | 6885 | hCoV-19/England/OXON-B01FD/2020\|EPI_ISL_478865\|202 | .................... | .................... | .................... | .................... | .................... | .................... | |
| D | 6886 | hCoV-19/England/OXON-B0209/2020\|EPI_ISL_478866\|202 | .................... | .................... | .................... | .................... | .................... | .................... | |
| D | 6887 | hCoV-19/England/OXON-B0236/2020\|EPI_ISL_478868\|202 | .................... | .................... | .................... | .................... | .................... | .................... | |
| D | 6888 | hCoV-19/England/OXON-B0272/2020\|EPI_ISL_478869\|202 | .................... | .................... | .................... | .................... | .................... | .................... | |
| D | 6889 | hCoV-19/England/OXON-B0281/2020\|EPI_ISL_478870\|202 | .................... | .................... | .................... | .................... | .................... | .................... | |
| D | 6890 | hCoV-19/England/OXON-B02DC/2020\|EPI_ISL_478872\|202 | .................... | .................... | .................... | .................... | .................... | .................... | |
| D | 6891 | hCoV-19/England/OXON-B02FA/2020\|EPI_ISL_478873\|202 | .................... | .................... | .................... | .................... | .................... | .................... | |
| D | 6892 | hCoV-19/England/OXON-B0315/2020\|EPI_ISL_478874\|202 | .................... | .................... | .................... | .................... | .................... | .................... | |
| D | 6893 | hCoV-19/England/OXON-B0351/2020\|EPI_ISL_478875\|202 | .................... | .................... | .................... | .................... | .................... | .................... | |
| D | 6894 | hCoV-19/England/OXON-B038E/2020\|EPI_ISL_478876\|202 | .................... | .................... | .................... | .................... | .................... | .................... | |
| D | 6895 | hCoV-19/England/OXON-B03BB/2020\|EPI_ISL_478877\|202 | .................... | .................... | .................... | .................... | .................... | .................... | |
| D | 6896 | hCoV-19/England/OXON-B0403/2020\|EPI_ISL_478878\|202 | .................... | .................... | .................... | .................... | .................... | .................... | |
| D | 6897 | hCoV-19/England/OXON-B0412/2020\|EPI_ISL_478879\|202 | .................... | .................... | .................... | .................... | .................... | .................... | |
| D | 6898 | hCoV-19/England/OXON-B0430/2020\|EPI_ISL_478880\|202 | .................... | .................... | .................... | .................... | .................... | .................... | |
| D | 6899 | hCoV-19/England/OXON-B044F/2020\|EPI_ISL_478881\|202 | .................... | .................... | .................... | .................... | .................... | .................... | |
| D | 6900 | hCoV-19/England/OXON-B049A/2020\|EPI_ISL_478882\|202 | .................... | .................... | .................... | .................... | .................... | .................... | |
| D | 6901 | hCoV-19/England/OXON-B04A9/2020\|EPI_ISL_478883\|202 | .................... | .................... | .................... | .................... | .................... | .................... | |
| D | 6902 | hCoV-19/England/OXON-B052E/2020\|EPI_ISL_478885\|202 | .................... | .................... | .................... | .................... | .................... | .................... | |
| D | 6903 | hCoV-19/England/OXON-B053D/2020\|EPI_ISL_478886\|202 | .................... | .................... | .................... | .................... | .................... | .................... | |
| D | 6904 | hCoV-19/England/OXON-B055B/2020\|EPI_ISL_478887\|202 | .................... | .................... | .................... | .................... | .................... | .................... | |

| | | | | | | | | |
|---|---|---|---|---|---|---|---|---|
| D | 6905 hCoV-19/England/OXON-B0579/2020\|EPI_ISL_478888\|202 | ···················· | ···················· | ···················· | ···················· | ···················· | ···················· | |
| D | 6906 hCoV-19/England/OXON-B0597/2020\|EPI_ISL_478889\|202 | ···················· | ···················· | ···················· | ···················· | ···················· | ···················· | |
| D | 6907 hCoV-19/England/OXON-B060D/2020\|EPI_ISL_478891\|202 | ···················· | ···················· | ···················· | ···················· | ···················· | ···················· | |
| D | 6908 hCoV-19/England/OXON-B062B/2020\|EPI_ISL_478892\|202 | ···················· | ···················· | ···················· | ···················· | ···················· | ···················· | |
| D | 6909 hCoV-19/England/OXON-B063A/2020\|EPI_ISL_478893\|202 | ···················· | ···················· | ···················· | ···················· | ···················· | ···················· | |
| D | 6910 hCoV-19/England/OXON-B0649/2020\|EPI_ISL_478894\|202 | ···················· | ···················· | ···················· | ···················· | ···················· | ···················· | |
| D | 6911 hCoV-19/England/OXON-B0658/2020\|EPI_ISL_478895\|202 | ···················· | ···················· | ···················· | ···················· | ···················· | ···················· | |
| D | 6912 hCoV-19/England/OXON-B0667/2020\|EPI_ISL_478896\|202 | ···················· | ···················· | ···················· | ···················· | ···················· | ···················· | |
| D | 6913 hCoV-19/Belgium/HN-0317324/2020\|EPI_ISL_462178\|202 | ···················· | ···················· | ···················· | ···················· | ···················· | ···················· | |
| D | 6914 hCoV-19/Belgium/WVM-0317325/2020\|EPI_ISL_462179\|20 | ···················· | ···················· | ···················· | ···················· | ···················· | ···················· | |
| D | 6915 hCoV-19/Belgium/JBE-0317326/2020\|EPI_ISL_462180\|20 | ···················· | ···················· | ···················· | ···················· | ···················· | ···················· | |
| D | 6916 hCoV-19/Belgium/HMM-0317327/2020\|EPI_ISL_462181\|20 | ···················· | ···················· | ···················· | ···················· | ···················· | ···················· | |
| D | 6917 hCoV-19/Belgium/CGR-0330328/2020\|EPI_ISL_462182\|20 | ···················· | ···················· | ···················· | ···················· | ···················· | ···················· | |
| D | 6918 hCoV-19/Belgium/VBWF-0330329/2020\|EPI_ISL_462183\|2 | ···················· | ···················· | ···················· | ···················· | ···················· | ···················· | |
| D | 6919 hCoV-19/Belgium/HMJ-0330330/2020\|EPI_ISL_462184\|20 | ···················· | ···················· | ···················· | ···················· | ···················· | ···················· | |
| D | 6920 hCoV-19/Belgium/SRL-0330331/2020\|EPI_ISL_462185\|20 | ···················· | ···················· | ···················· | ···················· | ···················· | ···················· | |
| D | 6921 hCoV-19/Belgium/CL-0330332/2020\|EPI_ISL_462186\|202 | ···················· | ···················· | ···················· | ···················· | ···················· | ···················· | |
| D | 6922 hCoV-19/Belgium/DWJ-0324333/2020\|EPI_ISL_462187\|20 | ···················· | ···················· | ···················· | ···················· | ···················· | ···················· | |
| D | 6923 hCoV-19/Belgium/CGP-0325334/2020\|EPI_ISL_462188\|20 | ···················· | ···················· | ···················· | ···················· | ···················· | ···················· | |
| D | 6924 hCoV-19/Belgium/BA-0330335/2020\|EPI_ISL_462189\|202 | ···················· | ···················· | ···················· | ···················· | ···················· | ···················· | |
| D | 6925 hCoV-19/Belgium/DGL-0326336/2020\|EPI_ISL_462190\|20 | ···················· | ···················· | ···················· | ···················· | ···················· | ···················· | |
| D | 6926 hCoV-19/Belgium/VMC-0325337/2020\|EPI_ISL_462191\|20 | ···················· | ···················· | ···················· | ···················· | ···················· | ···················· | |
| D | 6927 hCoV-19/Belgium/HJ-0328338/2020\|EPI_ISL_462192\|202 | ···················· | ···················· | ···················· | ···················· | ···················· | ···················· | |
| D | 6928 hCoV-19/Belgium/LAM-0325339/2020\|EPI_ISL_462193\|20 | ···················· | ···················· | ···················· | ···················· | ···················· | ···················· | |
| D | 6929 hCoV-19/Belgium/DBWC-0325340/2020\|EPI_ISL_462194\|2 | ···················· | ···················· | ···················· | ···················· | ···················· | ···················· | |
| D | 6930 hCoV-19/Belgium/LL-0328341/2020\|EPI_ISL_462195\|202 | ···················· | ···················· | ···················· | ···················· | ···················· | ···················· | |
| D | 6931 hCoV-19/Belgium/AM-0328342/2020\|EPI_ISL_462196\|202 | ···················· | ···················· | ···················· | ···················· | ···················· | ···················· | |
| D | 6932 hCoV-19/Belgium/NA-0328342/2020\|EPI_ISL_462197\|202 | ···················· | ···················· | ···················· | ···················· | ···················· | ···················· | |
| D | 6933 hCoV-19/Belgium/MR-0327344/2020\|EPI_ISL_462198\|202 | ···················· | ···················· | ···················· | ···················· | ···················· | ···················· | |
| D | 6934 hCoV-19/Belgium/CF-0327345/2020\|EPI_ISL_462199\|202 | ···················· | ···················· | ···················· | ···················· | ···················· | ···················· | |
| D | 6935 hCoV-19/England/20088028304/2020\|EPI_ISL_464180\|20 | ···················· | ···················· | ···················· | ···················· | ···················· | ···················· | |
| D | 6936 hCoV-19/England/20092000704/2020\|EPI_ISL_464181\|20 | ···················· | ···················· | ···················· | ···················· | ···················· | N··················· | |
| D | 6937 hCoV-19/England/20092000804/2020\|EPI_ISL_464182\|20 | ···················· | ···················· | ···················· | ···················· | ···················· | ···················· | |
| D | 6938 hCoV-19/England/20092096704/2020\|EPI_ISL_464183\|20 | ···················· | ···················· | ···················· | ···················· | K··················· | ···················· | |
| D | 6939 hCoV-19/England/20094007004/2020\|EPI_ISL_464184\|20 | ···················· | ···················· | ···················· | ···················· | ···················· | ···················· | |
| D | 6940 hCoV-19/England/20094061404/2020\|EPI_ISL_464185\|20 | ···················· | ···················· | ···················· | ···················· | ···················· | ···················· | |
| D | 6941 hCoV-19/England/20094061504/2020\|EPI_ISL_464186\|20 | ···················· | ···················· | ···················· | ···················· | ···················· | ···················· | |
| D | 6942 hCoV-19/England/20096051004/2020\|EPI_ISL_464187\|20 | ···················· | ···················· | ···················· | ···················· | ···················· | ···················· | |
| D | 6944 hCoV-19/England/20096101804/2020\|EPI_ISL_464189\|20 | ···················· | ···················· | ···················· | ···················· | ···················· | ···················· | |
| D | 6945 hCoV-19/England/20096115304/2020\|EPI_ISL_464190\|20 | ···················· | ···················· | ···················· | ···················· | ···················· | ···················· | |
| D | 6946 hCoV-19/England/20098017104/2020\|EPI_ISL_464192\|20 | ···················· | ···················· | ···················· | ···················· | ···················· | ···················· | |
| D | 6947 hCoV-19/England/20098017204/2020\|EPI_ISL_464193\|20 | ···················· | ···················· | ···················· | ···················· | ···················· | ·············N······ | |
| D | 6948 hCoV-19/England/20098017504/2020\|EPI_ISL_464194\|20 | ···················· | ···················· | ···················· | ···················· | ···················· | ···················· | |
| D | 6949 hCoV-19/England/20098017604/2020\|EPI_ISL_464195\|20 | ···················· | ···················· | ···················· | ···················· | ···················· | ···················· | |
| D | 6950 hCoV-19/England/20098017704/2020\|EPI_ISL_464196\|20 | ···················· | ···················· | ···················· | ···················· | ···················· | ···················· | |
| D | 6951 hCoV-19/England/20098017804/2020\|EPI_ISL_464197\|20 | ···················· | ···················· | ···················· | ···················· | ···················· | ···················· | |
| D | 6952 hCoV-19/England/20099000104/2020\|EPI_ISL_464198\|20 | ···················· | ···················· | ···················· | ···················· | ···················· | ···················· | |
| D | 6953 hCoV-19/England/20099000304/2020\|EPI_ISL_464199\|20 | ···················· | ···················· | ···················· | ···················· | ···················· | ···················· | |
| D | 6959 hCoV-19/Spain/IB-IBV-2501/2020\|EPI_ISL_469008\|2020 | ···················· | ···················· | ···················· | ···················· | ·NNN················ | ···················· | 1 |
| D | 6965 hCoV-19/Spain/IB-IBV-2590/2020\|EPI_ISL_469015\|2020 | ···················· | ···················· | ···················· | ···················· | ···················· | ···················· | |
| D | 6966 hCoV-19/Morocco/RMPS-01/2020\|EPI_ISL_469017\|2020-0 | ···················· | ···················· | ···················· | ···················· | ···················· | ···················· | |
| D | 6967 hCoV-19/Italy/APU-UniMI-809/2020\|EPI_ISL_469020\|20 | ···················· | ···················· | ···················· | ···················· | ···················· | ···················· | |
| D | 6968 hCoV-19/Italy/APU-UniMI-847/2020\|EPI_ISL_469022\|20 | ···················· | ···················· | ···················· | ···················· | ···················· | ···················· | |
| D | 6969 hCoV-19/Italy/APU-UniMI-857/2020\|EPI_ISL_469023\|20 | ···················· | ···················· | ···················· | ···················· | ···················· | ···················· | |
| D | 6970 hCoV-19/India/GJ-GBRC184/2020\|EPI_ISL_469024\|2020- | ···················· | ·A·················· | ···················· | ···················· | ···················· | ···A················ | |
| D | 6971 hCoV-19/India/GJ-GBRC185b/2020\|EPI_ISL_469026\|2020 | ···················· | ···················· | ···················· | ···················· | ···················· | ···················· | |
| D | 6972 hCoV-19/India/GJ-GBRC186a/2020\|EPI_ISL_469027\|2020 | ···················· | ···················· | ···················· | ···················· | ···················· | ···A················ | |
| D | 6973 hCoV-19/India/GJ-GBRC186b/2020\|EPI_ISL_469028\|2020 | ···················· | ·A·················· | ···················· | ···················· | ···················· | ···················· | |
| D | 6974 hCoV-19/India/GJ-GBRC187a/2020\|EPI_ISL_469029\|2020 | ···················· | ···················· | ···················· | ···················· | ···················· | ···················· | |
| D | 6975 hCoV-19/India/GJ-GBRC187b/2020\|EPI_ISL_469030\|2020 | ···················· | ···················· | ···················· | ···················· | ···················· | ···················· | |
| D | 6976 hCoV-19/India/GJ-GBRC188/2020\|EPI_ISL_469031\|2020- | ···················· | ···················· | ···················· | ···················· | ···················· | ···················· | |
| D | 6977 hCoV-19/India/GJ-GBRC189/2020\|EPI_ISL_469032\|2020- | ···················· | ···················· | ···················· | ···················· | ···················· | ···················· | |
| D | 6978 hCoV-19/India/GJ-GBRC190/2020\|EPI_ISL_469033\|2020- | ···················· | ···················· | ···················· | ···················· | ···················· | ···················· | |
| D | 6979 hCoV-19/India/GJ-GBRC191a/2020\|EPI_ISL_469034\|2020 | ···················· | ···················· | ···················· | ···················· | ···················· | ···················· | |
| D | 6980 hCoV-19/India/GJ-GBRC191b/2020\|EPI_ISL_469035\|2020 | ···················· | ···················· | ···················· | ···················· | ···················· | ···················· | |
| D | 6981 hCoV-19/India/GJ-GBRC192/2020\|EPI_ISL_469036\|2020- | ···················· | ···················· | ···················· | ···················· | ···················· | ···················· | |
| D | 6982 hCoV-19/India/GJ-GBRC193a/2020\|EPI_ISL_469037\|2020 | ···················· | ···················· | ···················· | ···················· | ···················· | ···················· | |
| D | 6983 hCoV-19/India/GJ-GBRC193b/2020\|EPI_ISL_469038\|2020 | ···················· | ···················· | ···················· | ···················· | ···················· | ···A················ | |
| D | 6984 hCoV-19/India/GJ-GBRC195a/2020\|EPI_ISL_469039\|2020 | ···················· | ···················· | ···················· | ···················· | ···················· | ···················· | |
| D | 6985 hCoV-19/India/GJ-GBRC195b/2020\|EPI_ISL_469040\|2020 | ···················· | ···················· | ···················· | ···················· | ···················· | ···················· | |
| D | 6986 hCoV-19/India/GJ-GBRC196/2020\|EPI_ISL_469041\|2020- | ···················· | ···················· | ···················· | ···················· | ···················· | ···················· | |
| D | 6987 hCoV-19/India/GJ-GBRC197/2020\|EPI_ISL_469042\|2020- | ···················· | ···················· | ···················· | ···················· | ···················· | ···················· | |
| D | 6988 hCoV-19/India/GJ-GBRC198/2020\|EPI_ISL_469043\|2020- | ···················· | ···················· | ···················· | ···················· | ···················· | ···················· | |
| D | 6989 hCoV-19/India/GJ-GBRC199/2020\|EPI_ISL_469044\|2020- | ···················· | ·A·················· | ···················· | ···················· | ···················· | ···A················ | |
| D | 6990 hCoV-19/India/GJ-GBRC200/2020\|EPI_ISL_469045\|2020- | ···················· | ·A·················· | ···················· | ···················· | ···················· | ···A················ | |
| D | 6991 hCoV-19/India/GJ-GBRC201a/2020\|EPI_ISL_469046\|2020 | ···················· | ·A·················· | ···················· | ···················· | ···················· | ···A················ | |
| D | 6992 hCoV-19/India/GJ-GBRC201b/2020\|EPI_ISL_469047\|2020 | ···················· | ·A·················· | ···················· | ···················· | ···················· | ···A················ | |
| D | 6993 hCoV-19/India/GJ-GBRC202/2020\|EPI_ISL_469048\|2020- | ···················· | ···················· | ···················· | ···················· | ···················· | ···················· | |

| | | | | | | | | |
|---|---|---|---|---|---|---|---|---|
| D | 6994 hCoV-19/Morocco/RMPS-02/2020\|EPI_ISL_469049\|2020-0 | ···················· | ···················· | ····················· | ···················· | ···················· | ···················· | |
| D | 6995 hCoV-19/Morocco/RMPS-03/2020\|EPI_ISL_469051\|2020-0 | ···················· | ···················· | ····················· | ···················· | ···················· | ···················· | |
| D | 6996 hCoV-19/Morocco/RMPS-04/2020\|EPI_ISL_469052\|2020-0 | ···················· | ···················· | ····················· | ···················· | ···················· | ···················· | |
| D | 6997 hCoV-19/Morocco/RMPS-05/2020\|EPI_ISL_469053\|2020-0 | ···················· | ···················· | ····················· | ···················· | ···················· | ···················· | |
| D | 6998 hCoV-19/Morocco/RMPS-06/2020\|EPI_ISL_469054\|2020-0 | ···················· | ···················· | ····················· | ···················· | ···················· | ···················· | |
| D | 6999 hCoV-19/Sweden/20-14262/2020\|EPI_ISL_469055\|2020-0 | ···················· | ···················· | ····················· | ···················· | ···················· | ···················· | |
| D | 7000 hCoV-19/Sweden/20-14317/2020\|EPI_ISL_469056\|2020-0 | ···················· | ···················· | ····················· | ···················· | ···················· | ···················· | |
| D | 7001 hCoV-19/Sweden/20-14318/2020\|EPI_ISL_469057\|2020-0 | ···················· | ···················· | ····················· | ···················· | ···················· | ···················· | |
| D | 7002 hCoV-19/Sweden/20-14377/2020\|EPI_ISL_469058\|2020-0 | ···················· | ···················· | ····················· | ···················· | ···················· | ···················· | |
| D | 7003 hCoV-19/Sweden/20-14647/2020\|EPI_ISL_469059\|2020-0 | ···················· | ···················· | ····················· | ···················· | ···················· | ···················· | |
| D | 7004 hCoV-19/Sweden/20-14696/2020\|EPI_ISL_469060\|2020-0 | ···················· | ···················· | ····················· | ···················· | ···················· | ···················· | |
| D | 7005 hCoV-19/Sweden/20-14697/2020\|EPI_ISL_469061\|2020-0 | ···················· | ···················· | ····················· | ···················· | ···················· | ···················· | |
| D | 7006 hCoV-19/Sweden/20-14785/2020\|EPI_ISL_469062\|2020-0 | ···················· | ···················· | ····················· | ···················· | ···················· | ···················· | |
| D | 7007 hCoV-19/Sweden/20-14827/2020\|EPI_ISL_469063\|2020-0 | ···················· | ···················· | ····················· | ···················· | ···················· | ···················· | |
| D | 7008 hCoV-19/Sweden/20-19008/2020\|EPI_ISL_469064\|2020-0 | ···················· | ···················· | ··A·················· | ···················· | ···················· | ···················· | |
| D | 7009 hCoV-19/Sweden/20-19192/2020\|EPI_ISL_469065\|2020-0 | ···················· | ···················· | ····················· | ···················· | ···················· | ···················· | |
| D | 7010 hCoV-19/Sweden/20-51350/2020\|EPI_ISL_469066\|2020-0 | ···················· | ···················· | ····················· | ···················· | ···················· | ···················· | |
| D | 7011 hCoV-19/Sweden/20-51598/2020\|EPI_ISL_469067\|2020-0 | ···················· | ···················· | ····················· | ···················· | ···················· | ···················· | |
| D | 7012 hCoV-19/Sweden/20-51618/2020\|EPI_ISL_469068\|2020-0 | ···················· | ···················· | ····················· | ···················· | ···················· | ···················· | |
| D | 7013 hCoV-19/Sweden/20-51678/2020\|EPI_ISL_469069\|2020-0 | ···················· | ···················· | ····················· | ···················· | ···················· | ···················· | |
| D | 7014 hCoV-19/Scotland/GCVR-17219C/2020\|EPI_ISL_470000\|2 | ···················· | ···················· | ····················· | ···················· | ···················· | ···················· | |
| D | 7015 hCoV-19/Scotland/GCVR-17235A/2020\|EPI_ISL_470001\|2 | ···················· | ···················A | ····················· | ···················· | ···················· | ···················· | 1 |
| D | 7016 hCoV-19/Scotland/GCVR-172053/2020\|EPI_ISL_470005\|2 | ···················· | ···················· | ····················· | ···················· | ···················· | ···················· | |
| D | 7017 hCoV-19/Scotland/GCVR-171616/2020\|EPI_ISL_470006\|2 | ···················· | ···················· | ····················· | ···················· | ···················· | ···················· | |
| D | 7018 hCoV-19/Scotland/GCVR-1724A2/2020\|EPI_ISL_470007\|2 | ···················· | ···················A | ····················· | ···················· | ···················· | ···················· | 1 |
| D | 7019 hCoV-19/Scotland/GCVR-17234B/2020\|EPI_ISL_470009\|2 | ···················· | ···················· | ····················· | ···················· | ···················· | ···················· | |
| D | 7020 hCoV-19/Scotland/GCVR-171F2A/2020\|EPI_ISL_470010\|2 | ···················· | ···················· | ····················· | ···················· | ···················· | ···················· | |
| D | 7021 hCoV-19/Northern Ireland/NIRE-FB897/2020\|EPI_ISL_4 | ···················· | ···················· | ····················· | ···················· | ···················· | ···················· | |
| D | 7022 hCoV-19/Northern Ireland/NIRE-FBB52/2020\|EPI_ISL_4 | ···················· | ···················· | ··········T·········· | ···················· | ···················· | ···················· | |
| D | 7023 hCoV-19/Northern Ireland/NIRE-FB080/2020\|EPI_ISL_4 | ···················· | ···················· | ····················· | ···················· | ···················· | ···················· | |
| D | 7024 hCoV-19/Northern Ireland/NIRE-FC19B/2020\|EPI_ISL_4 | ···················· | ···················· | ····················· | ···················· | ···················· | ···················· | |
| D | 7025 hCoV-19/Northern Ireland/NIRE-FAF2A/2020\|EPI_ISL_4 | ···················· | ···················· | ····················· | ···················· | ···················· | ···················· | |
| D | 7026 hCoV-19/Northern Ireland/NIRE-FC1E6/2020\|EPI_ISL_4 | ···················· | ···················· | ····················· | ···················· | ···················· | ···················· | |
| D | 7027 hCoV-19/Northern Ireland/NIRE-FB105/2020\|EPI_ISL_4 | ···················· | ···················· | ····················· | ···················· | ···················· | ···················· | |
| D | 7028 hCoV-19/Northern Ireland/NIRE-FC9C0/2020\|EPI_ISL_4 | ···················· | ···················· | ····················· | ···················· | ···················· | ···················· | |
| D | 7029 hCoV-19/Northern Ireland/NIRE-FC77B/2020\|EPI_ISL_4 | ···················· | ···················· | ····················· | ···················· | ···················· | ···················· | |
| D | 7030 hCoV-19/Northern Ireland/NIRE-FC0DA/2020\|EPI_ISL_4 | ···················· | ···················· | ····················· | ···················· | ···················· | ···················· | |
| D | 7031 hCoV-19/Northern Ireland/NIRE-FBCE6/2020\|EPI_ISL_4 | ···················· | ···················· | ····················· | ···················· | ···················· | ···················· | |
| D | 7032 hCoV-19/Northern Ireland/NIRE-FBA91/2020\|EPI_ISL_4 | ···················· | ···················· | ····················· | ···················· | ···················· | ···················· | |
| D | 7033 hCoV-19/Northern Ireland/NIRE-FCA09/2020\|EPI_ISL_4 | ···················· | ···················· | ····················· | ···················· | ···················· | ···················· | |
| D | 7034 hCoV-19/Northern Ireland/NIRE-FBC9B/2020\|EPI_ISL_4 | ···················· | ···················· | ····················· | ···················· | ···················· | ···················· | |
| D | 7035 hCoV-19/Northern Ireland/NIRE-FC5F9/2020\|EPI_ISL_4 | ···················· | ···················· | ····················· | ···················· | ···················· | ···················· | |
| D | 7036 hCoV-19/Northern Ireland/NIRE-FB132/2020\|EPI_ISL_4 | ···················· | ···················· | ····················· | ···················· | ···················· | ···················· | |
| D | 7037 hCoV-19/Northern Ireland/NIRE-FBDD4/2020\|EPI_ISL_4 | ···················· | ···················· | ····················· | ···················· | ···················· | ···················· | |
| D | 7038 hCoV-19/Northern Ireland/NIRE-FBA19/2020\|EPI_ISL_4 | ···················· | ···················· | ····················· | ···················· | ···················· | ···················· | |
| D | 7039 hCoV-19/Northern Ireland/NIRE-FBC6E/2020\|EPI_ISL_4 | ···················· | ···················· | ····················· | ···················· | ···················· | ···················· | |
| D | 7040 hCoV-19/Northern Ireland/NIRE-FC526/2020\|EPI_ISL_4 | ···················· | ···················· | ····················· | ···················· | ···················· | ···················· | |
| D | 7041 hCoV-19/Northern Ireland/NIRE-FBDF2/2020\|EPI_ISL_4 | ···················· | ···················· | ····················· | ···················· | ···················· | ···················· | |
| D | 7042 hCoV-19/Northern Ireland/NIRE-FCC21/2020\|EPI_ISL_4 | ···················· | ···················· | ····················· | ···················· | ···················· | ···················· | |
| D | 7043 hCoV-19/Northern Ireland/NIRE-FBE95/2020\|EPI_ISL_4 | ···················· | ···················· | ····················· | ···················· | ···················· | ···················· | |
| D | 7044 hCoV-19/Northern Ireland/NIRE-FC0CB/2020\|EPI_ISL_4 | ···················· | ···················· | ····················· | ···················· | ···················· | ···················· | |
| D | 7045 hCoV-19/USA/MI-MDHHS-SC20879/2020\|EPI_ISL_471900\|2 | ···················· | ··C················· | ····················· | ···················· | ···················· | ···················· | |
| D | 7046 hCoV-19/USA/MI-MDHHS-SC20880/2020\|EPI_ISL_471901\|2 | ···················· | ···················· | ····················· | ···················· | ···················· | ···················· | |
| D | 7047 hCoV-19/USA/MI-MDHHS-SC20881/2020\|EPI_ISL_471902\|2 | ···················· | ···················· | ····················· | ···················· | ···················· | ···················· | |
| D | 7048 hCoV-19/USA/MI-MDHHS-SC20882/2020\|EPI_ISL_471903\|2 | ···················· | ···················· | ····················· | ···················· | ···················· | ···················· | |
| D | 7049 hCoV-19/USA/MI-MDHHS-SC20883/2020\|EPI_ISL_471904\|2 | ···················· | ···················· | ····················· | ···················· | ···················· | ···················· | |
| D | 7050 hCoV-19/USA/MI-MDHHS-SC20884/2020\|EPI_ISL_471905\|2 | ···················· | ···················· | ····················· | ···················· | ···················· | ···················· | |
| D | 7051 hCoV-19/USA/MI-MDHHS-SC20885/2020\|EPI_ISL_471906\|2 | ···················· | ···················· | ····················· | ···················· | ···················· | ···················· | |
| D | 7052 hCoV-19/USA/MI-MDHHS-SC20886/2020\|EPI_ISL_471907\|2 | ···················· | ···················· | ····················· | ···················· | ···················· | ···················· | |
| D | 7053 hCoV-19/USA/MI-MDHHS-SC20887/2020\|EPI_ISL_471908\|2 | ···················· | ···················· | ····················· | ···················· | ···················· | ···················· | |
| D | 7054 hCoV-19/USA/MI-MDHHS-SC20888/2020\|EPI_ISL_471909\|2 | ···················· | ···················· | ····················· | ···················· | ···················· | ···················· | |
| D | 7055 hCoV-19/USA/MI-MDHHS-SC20889/2020\|EPI_ISL_471910\|2 | ···················· | ···················· | ····················· | ···················· | ···················· | ···················· | |
| D | 7056 hCoV-19/England/EXET-136CB1/2020\|EPI_ISL_471911\|20 | ···················· | ···················· | ····················· | ···················· | ···················· | ···················· | |
| D | 7057 hCoV-19/England/EXET-136CC0/2020\|EPI_ISL_471912\|20 | ···················· | ···················· | ····················· | ···················· | ···················· | ···················· | |
| D | 7058 hCoV-19/England/EXET-136CDF/2020\|EPI_ISL_471913\|20 | ···················· | ···················· | ····················· | ···················· | ···················· | ···················· | |
| D | 7059 hCoV-19/England/EXET-136CFD/2020\|EPI_ISL_471914\|20 | ···················· | ···················· | ····················· | ···················· | ···················· | ···················· | |
| D | 7060 hCoV-19/England/EXET-136D09/2020\|EPI_ISL_471915\|20 | ···················· | ···················· | ····················· | ···················· | ···················· | ···················· | |
| D | 7061 hCoV-19/England/EXET-136D18/2020\|EPI_ISL_471916\|20 | ···················· | ···················· | ····················· | ···················· | ···················· | ···················· | |
| D | 7062 hCoV-19/England/EXET-136D36/2020\|EPI_ISL_471917\|20 | ···················· | ···················· | ····················· | ···················· | ···················· | ···················· | |
| D | 7063 hCoV-19/England/EXET-136D45/2020\|EPI_ISL_471918\|20 | ···················· | ···················· | ····················· | ···················· | ···················· | ···················· | |
| D | 7064 hCoV-19/England/EXET-136D54/2020\|EPI_ISL_471919\|20 | ···················· | ···················· | ····················· | ···················· | ···················· | ···················· | |
| D | 7065 hCoV-19/Wales/PHWC-15CC88/2020\|EPI_ISL_472602\|2020 | ···················· | ···················· | ····················· | ···················· | ···················· | ···················· | |
| D | 7066 hCoV-19/Wales/PHWC-15CC97/2020\|EPI_ISL_472603\|2020 | ···················· | ···················· | ····················· | ···················· | ···················· | ···················· | |
| D | 7067 hCoV-19/Wales/PHWC-15CCB5/2020\|EPI_ISL_472605\|2020 | ···················· | ···················· | ····················· | ···················· | ···················· | ···················· | |
| D | 7068 hCoV-19/Wales/PHWC-15CCC4/2020\|EPI_ISL_472606\|2020 | ···················· | ···················· | ····················· | ···················· | ···················· | ···················· | |
| D | 7069 hCoV-19/Wales/PHWC-15CD67/2020\|EPI_ISL_472615\|2020 | ···················· | ···················· | ····················· | ···················· | ···················· | ···················· | |
| D | 7070 hCoV-19/Wales/PHWC-15CD94/2020\|EPI_ISL_472618\|2020 | ···················· | ···················· | ····················· | ···················· | ···················· | ···················· | |
| D | 7071 hCoV-19/Wales/PHWC-15CDEF/2020\|EPI_ISL_472622\|2020 | ···················· | ···················· | ····················· | ···················· | ···················· | ···················· | |

| | | | | | | | | |
|---|---|---|---|---|---|---|---|---|
| D | 7072 hCoV-19/Wales/PHWC-15CDFE/2020\|EPI_ISL_472623\|2020 | .................... | .................... | .................... | .................... | .................... | .................... | |
| D | 7073 hCoV-19/Wales/PHWC-15CE19/2020\|EPI_ISL_472625\|2020 | .................... | .................... | .................... | .................... | .................... | .................... | |
| D | 7074 hCoV-19/Wales/PHWC-15CE37/2020\|EPI_ISL_472626\|2020 | .................... | .................... | .................... | .................... | .................... | .................... | |
| D | 7075 hCoV-19/Wales/PHWC-15CEDD/2020\|EPI_ISL_472636\|2020 | .................... | .................... | .................... | .................... | .................... | .................... | |
| D | 7076 hCoV-19/Wales/PHWC-15CEEC/2020\|EPI_ISL_472637\|2020 | .................... | .................... | .................... | .................... | .................... | .................... | |
| D | 7077 hCoV-19/Wales/PHWC-15CF07/2020\|EPI_ISL_472639\|2020 | .................... | .................... | .................... | .................... | .................... | .................... | |
| D | 7078 hCoV-19/Wales/PHWC-15CFE9/2020\|EPI_ISL_472653\|2020 | .................... | .................... | .................... | .................... | .................... | .................... | |
| D | 7079 hCoV-19/Wales/PHWC-15D003/2020\|EPI_ISL_472655\|2020 | .................... | .................... | .................... | .................... | .................... | .................... | |
| D | 7080 hCoV-19/Wales/PHWC-15D09A/2020\|EPI_ISL_472664\|2020 | .................... | .................... | .................... | .................... | .................... | .................... | |
| D | 7081 hCoV-19/Wales/PHWC-15D0C7/2020\|EPI_ISL_472666\|2020 | .................... | .................... | .................... | .................... | .................... | .................... | |
| D | 7082 hCoV-19/Wales/PHWC-15D11F/2020\|EPI_ISL_472669\|2020 | .................... | .................... | .................... | .................... | .................... | .................... | |
| D | 7083 hCoV-19/Wales/PHWC-15D12E/2020\|EPI_ISL_472670\|2020 | .................... | .................... | .................... | .................... | .................... | .................... | |
| D | 7084 hCoV-19/Wales/PHWC-15D20D/2020\|EPI_ISL_472684\|2020 | .................... | .................... | .................... | .................... | .................... | .................... | |
| D | 7085 hCoV-19/Wales/PHWC-15D285/2020\|EPI_ISL_472692\|2020 | .................... | .................... | .................... | .................... | .................... | .................... | |
| D | 7086 hCoV-19/Wales/PHWC-15D294/2020\|EPI_ISL_472693\|2020 | .................... | .................... | .................... | .................... | .................... | .................... | |
| D | 7087 hCoV-19/Wales/PHWC-15D2FE/2020\|EPI_ISL_472699\|2020 | .................... | .................... | .................... | .................... | .................... | .................... | |
| D | 7088 hCoV-19/Scotland/EDB6171/2020\|EPI_ISL_473900\|2020- | .................... | .................... | .................... | .................... | .................... | .................... | |
| D | 7089 hCoV-19/Scotland/EDB6174/2020\|EPI_ISL_473901\|2020- | .................... | ...................A | .................... | .................... | .................... | .................... | 1 |
| D | 7090 hCoV-19/Scotland/EDB6176/2020\|EPI_ISL_473902\|2020- | .................... | ...................A | .................... | .................... | .................... | .................... | 1 |
| D | 7091 hCoV-19/Scotland/EDB6177/2020\|EPI_ISL_473903\|2020- | .................... | .................... | .................... | .................... | .................... | .................... | |
| D | 7092 hCoV-19/Scotland/EDB6178/2020\|EPI_ISL_473904\|2020- | .................... | ...................A | .................... | .................... | .................... | .................... | 1 |
| D | 7093 hCoV-19/Scotland/EDB6180/2020\|EPI_ISL_473905\|2020- | .................... | ...................A | .................... | .................... | .................... | .................... | 1 |
| D | 7094 hCoV-19/Scotland/EDB6184/2020\|EPI_ISL_473906\|2020- | .................... | ...................A | .................... | .................... | .................... | .................... | 1 |
| D | 7095 hCoV-19/Scotland/EDB6188/2020\|EPI_ISL_473907\|2020- | .................... | ...................A | .................... | .................... | .................... | .................... | 1 |
| D | 7096 hCoV-19/Scotland/EDB6192/2020\|EPI_ISL_473908\|2020- | .................... | ...................A | .................... | .................... | .................... | .................... | 1 |
| D | 7097 hCoV-19/Scotland/EDB6193/2020\|EPI_ISL_473909\|2020- | .................... | ...................A | .................... | .................... | .................... | .................... | 1 |
| D | 7098 hCoV-19/Scotland/EDB6194/2020\|EPI_ISL_473910\|2020- | .................... | .................... | .................... | .................... | .................... | .................... | |
| D | 7099 hCoV-19/Scotland/EDB6204/2020\|EPI_ISL_473911\|2020- | .................... | .................... | .................... | .................... | .................... | .................... | |
| D | 7100 hCoV-19/Scotland/EDB6205/2020\|EPI_ISL_473912\|2020- | .................... | ...................A | .................... | .................... | .................... | .................... | 1 |
| D | 7101 hCoV-19/Scotland/EDB6208/2020\|EPI_ISL_473913\|2020- | .................... | ...................A | .................... | .................... | .................... | .................... | 1 |
| D | 7102 hCoV-19/Scotland/EDB6209/2020\|EPI_ISL_473914\|2020- | .................... | .................... | .................... | .................... | .................... | .................... | |
| D | 7103 hCoV-19/Scotland/EDB6211/2020\|EPI_ISL_473915\|2020- | .................... | ...................A | .................... | .................... | .................... | .................... | 1 |
| D | 7104 hCoV-19/Scotland/EDB6212/2020\|EPI_ISL_473916\|2020- | .................... | .A.................. | .................... | .................... | .................... | .................... | |
| D | 7105 hCoV-19/Scotland/EDB6213/2020\|EPI_ISL_473917\|2020- | .................... | .................... | .................... | .................... | .................... | .................... | |
| D | 7106 hCoV-19/Scotland/EDB6214/2020\|EPI_ISL_473918\|2020- | .................... | ...................A | .................... | .................... | .................... | .................... | 1 |
| D | 7107 hCoV-19/Scotland/EDB6215/2020\|EPI_ISL_473919\|2020- | .................... | ...................A | .................... | .................... | .................... | .................... | 1 |
| D | 7108 hCoV-19/Scotland/EDB6222/2020\|EPI_ISL_473920\|2020- | .................... | ...................A | .................... | .................... | .................... | .................... | 1 |
| D | 7109 hCoV-19/Scotland/EDB6223/2020\|EPI_ISL_473921\|2020- | .................... | ...................A | .................... | .................... | .................... | .................... | 1 |
| D | 7110 hCoV-19/Scotland/EDB6224/2020\|EPI_ISL_473922\|2020- | .................... | ...................A | .................... | .................... | .................... | .................... | 1 |
| D | 7111 hCoV-19/Scotland/EDB6235/2020\|EPI_ISL_473924\|2020- | .................... | ...................A | .................... | .................... | .................... | .................... | 1 |
| D | 7112 hCoV-19/Scotland/EDB6243/2020\|EPI_ISL_473925\|2020- | .................... | ...................A | .................... | .................... | .................... | .................... | 1 |
| D | 7113 hCoV-19/Scotland/EDB6244/2020\|EPI_ISL_473926\|2020- | .................... | ...................A | .................... | .................... | .................... | .................... | 1 |
| D | 7114 hCoV-19/Scotland/EDB6245/2020\|EPI_ISL_473927\|2020- | .................... | ...................A | .................... | .................... | .................... | .................... | 1 |
| D | 7115 hCoV-19/Scotland/EDB6246/2020\|EPI_ISL_473928\|2020- | .................... | ...................A | .................... | .................... | .................... | .................... | 1 |
| D | 7116 hCoV-19/Scotland/EDB6247/2020\|EPI_ISL_473929\|2020- | .................... | ...................A | .................... | .................... | .................... | .................... | 1 |
| D | 7117 hCoV-19/Scotland/EDB6248/2020\|EPI_ISL_473930\|2020- | .................... | ...................A | .................... | .................... | .................... | .................... | 1 |
| D | 7118 hCoV-19/Scotland/EDB6251/2020\|EPI_ISL_473931\|2020- | .................... | ...................A | .................... | .................... | .................... | .................... | 1 |
| D | 7119 hCoV-19/Scotland/EDB6254/2020\|EPI_ISL_473932\|2020- | .................... | .................... | .................... | .................... | .................... | .................... | |
| D | 7120 hCoV-19/Scotland/EDB6259/2020\|EPI_ISL_473933\|2020- | .................... | .................... | .................... | .................... | .................... | .................... | |
| D | 7121 hCoV-19/Scotland/EDB6319/2020\|EPI_ISL_473934\|2020- | .................... | .A.................. | .................... | .................... | .................... | .................... | |
| D | 7122 hCoV-19/Scotland/EDB6322/2020\|EPI_ISL_473935\|2020- | .................... | .................... | .................... | .................... | .................... | .................... | |
| D | 7123 hCoV-19/Scotland/EDB6324/2020\|EPI_ISL_473936\|2020- | .................... | .................... | .................... | .................... | .................... | .................... | |
| D | 7124 hCoV-19/Scotland/EDB6328/2020\|EPI_ISL_473938\|2020- | .................... | .................... | .................... | .................... | .................... | .................... | |
| D | 7125 hCoV-19/Scotland/EDB6333/2020\|EPI_ISL_473940\|2020- | .................... | .................... | .................... | .................... | .................... | .................... | |
| D | 7126 hCoV-19/Scotland/EDB6356/2020\|EPI_ISL_473946\|2020- | .................... | .................... | .................... | .................... | .................... | .................... | |
| D | 7127 hCoV-19/Scotland/EDB6361/2020\|EPI_ISL_473947\|2020- | .................... | .................... | .................... | .................... | .................... | .................... | |
| D | 7128 hCoV-19/Scotland/EDB6369/2020\|EPI_ISL_473948\|2020- | .................... | .................... | .................... | .................... | .................... | .................... | |
| D | 7129 hCoV-19/Scotland/EDB6383/2020\|EPI_ISL_473949\|2020- | .................... | .................... | .................... | .................... | .................... | .................... | |
| D | 7130 hCoV-19/Scotland/EDB6391/2020\|EPI_ISL_473952\|2020- | .................... | .................... | .................... | .................... | .................... | .................... | |
| D | 7131 hCoV-19/Scotland/EDB6392/2020\|EPI_ISL_473953\|2020- | .................... | .................... | .................... | .................... | .................... | .................... | |
| D | 7132 hCoV-19/Scotland/EDB6394/2020\|EPI_ISL_473954\|2020- | ...........T........ | .................... | .................... | .................... | .................... | .................... | |
| D | 7133 hCoV-19/Wales/PHWC-16351A/2020\|EPI_ISL_473958\|2020 | .................... | .................... | .................... | .................... | .................... | .................... | |
| D | 7134 hCoV-19/Wales/PHWC-163538/2020\|EPI_ISL_473959\|2020 | .................... | .................... | .................... | .................... | .................... | .................... | |
| D | 7135 hCoV-19/Wales/PHWC-163547/2020\|EPI_ISL_473960\|2020 | .................... | .................... | .................... | .................... | .................... | .................... | |
| D | 7137 hCoV-19/Wales/PHWC-163565/2020\|EPI_ISL_473962\|2020 | .................... | .................... | .................... | .................... | .................... | .................... | |
| D | 7138 hCoV-19/Wales/PHWC-163592/2020\|EPI_ISL_473965\|2020 | .................... | .................... | .................... | .................... | .................... | .................... | |
| D | 7139 hCoV-19/Wales/PHWC-1635A1/2020\|EPI_ISL_473966\|2020 | .................... | .................... | .................... | .................... | .................... | .................... | |
| D | 7140 hCoV-19/Wales/PHWC-1635B0/2020\|EPI_ISL_473967\|2020 | .................... | .................... | .................... | .................... | .................... | .................... | |
| D | 7141 hCoV-19/Wales/PHWC-1635DE/2020\|EPI_ISL_473969\|2020 | .................... | .................... | .................... | .................... | .................... | .................... | |
| D | 7142 hCoV-19/Wales/PHWC-1635FC/2020\|EPI_ISL_473970\|2020 | .................... | .................... | .................... | .................... | .................... | .................... | |
| D | 7143 hCoV-19/Wales/PHWC-163608/2020\|EPI_ISL_473971\|2020 | .................... | .................... | .................... | .................... | .................... | .................... | |
| D | 7144 hCoV-19/Wales/PHWC-163644/2020\|EPI_ISL_473975\|2020 | .................... | .................... | .................... | .................... | .................... | .................... | |
| D | 7145 hCoV-19/Wales/PHWC-163653/2020\|EPI_ISL_473976\|2020 | .................... | .................... | .................... | .................... | .................... | .................... | |
| D | 7146 hCoV-19/Wales/PHWC-163662/2020\|EPI_ISL_473977\|2020 | .................... | .................... | .................... | .................... | .................... | .................... | |
| D | 7147 hCoV-19/Wales/PHWC-163680/2020\|EPI_ISL_473978\|2020 | .................... | .................... | .................... | .................... | .................... | .................... | |
| D | 7148 hCoV-19/Wales/PHWC-1636CC/2020\|EPI_ISL_473981\|2020 | .................... | .................... | .................... | .................... | .................... | .................... | |
| D | 7149 hCoV-19/Wales/PHWC-1636EA/2020\|EPI_ISL_473983\|2020 | .................... | .................... | .................... | .................... | .................... | .................... | |
| D | 7150 hCoV-19/Wales/PHWC-1636F9/2020\|EPI_ISL_473984\|2020 | .................... | .................... | .................... | .................... | .................... | .................... | |

| | | | | | | | | |
|---|---|---|---|---|---|---|---|---|
| D | 7151 | hCoV-19/Wales/PHWC-163714/2020\|EPI_ISL_473986\|2020 | ···················· | ···················· | ····················· | ···················· | ···················· | ···················· |
| D | 7152 | hCoV-19/Wales/PHWC-163732/2020\|EPI_ISL_473988\|2020 | ···················· | ···················· | ····················· | ···················· | ···················· | ···················· |
| D | 7153 | hCoV-19/Wales/PHWC-16376F/2020\|EPI_ISL_473990\|2020 | ···················· | ···················· | ····················· | ···················· | ···················· | ···················· |
| D | 7154 | hCoV-19/Wales/PHWC-16377E/2020\|EPI_ISL_473991\|2020 | ···················· | ···················· | ····················· | ···················· | ···················· | ···················· |
| D | 7155 | hCoV-19/Wales/PHWC-16378D/2020\|EPI_ISL_473992\|2020 | ···················· | ···················· | ····················· | ···················· | ···················· | ···················· |
| D | 7156 | hCoV-19/Wales/PHWC-16379C/2020\|EPI_ISL_473993\|2020 | ···················· | ···················· | ····················· | ···················· | ···················· | ···················· |
| D | 7157 | hCoV-19/Wales/PHWC-1637AB/2020\|EPI_ISL_473994\|2020 | ···················· | ···················· | ····················· | ···················· | ···················· | ···················· |
| D | 7158 | hCoV-19/Wales/PHWC-1637E7/2020\|EPI_ISL_473998\|2020 | ···················· | ···················· | ····················· | ···················· | ···················· | ···················· |
| D | 7159 | hCoV-19/Wales/PHWC-164458/2020\|EPI_ISL_474179\|2020 | ···················· | ···················· | ····················· | ···················· | ···················· | ···················· |
| D | 7160 | hCoV-19/Wales/PHWC-164476/2020\|EPI_ISL_474180\|2020 | ···················· | ···················· | ····················· | ···················· | ···················· | ···················· |
| D | 7161 | hCoV-19/Wales/PHWC-164485/2020\|EPI_ISL_474181\|2020 | ···················· | ···················· | ····················· | ···················· | ···················· | ···················· |
| D | 7162 | hCoV-19/Wales/PHWC-164494/2020\|EPI_ISL_474182\|2020 | ···················· | ···················· | ····················· | ···················· | ···················· | ···················· |
| D | 7163 | hCoV-19/Wales/PHWC-1644A3/2020\|EPI_ISL_474183\|2020 | ···················· | ···················· | ····················· | ···················· | ···················· | ···················· |
| D | 7164 | hCoV-19/Wales/PHWC-1644C1/2020\|EPI_ISL_474184\|2020 | ···················· | ···················· | ····················· | ···················· | ···················· | ···················· |
| D | 7165 | hCoV-19/Wales/PHWC-1644FE/2020\|EPI_ISL_474187\|2020 | ···················· | ···················· | ····················· | ···················· | ···················· | ···················· |
| D | 7166 | hCoV-19/Wales/PHWC-164519/2020\|EPI_ISL_474189\|2020 | ···················· | ·A·················· | ····················· | ···················· | ···················· | ···················· |
| D | 7167 | hCoV-19/Wales/PHWC-164528/2020\|EPI_ISL_474190\|2020 | ···················· | ···················· | ····················· | ···················· | ···················· | ···················· |
| D | 7168 | hCoV-19/Wales/PHWC-1645A0/2020\|EPI_ISL_474198\|2020 | ···················· | ···················· | ····················· | ···················· | ···················· | ···················· |
| D | 7169 | hCoV-19/Russia/CRIE160583/2020\|EPI_ISL_462149\|2020 | ···················· | ···················· | ····················· | ···················· | ···················· | ···················· |
| D | 7170 | hCoV-19/Russia/CRIE162784/2020\|EPI_ISL_462150\|2020 | ···················· | ···················· | ····················· | ···················· | ···················· | ···················· |
| D | 7171 | hCoV-19/Belgium/rega-0423297/2020\|EPI_ISL_462151\|2 | ···················· | ···················· | ····················· | ···················· | ···················· | ···················· |
| D | 7172 | hCoV-19/Belgium/rega-0423298/2020\|EPI_ISL_462152\|2 | ···················· | ···················· | ····················· | ···················· | ···················· | ···················· |
| D | 7173 | hCoV-19/Belgium/rega-0423299/2020\|EPI_ISL_462153\|2 | ···················· | ···················· | ····················· | ···················· | ···················· | ···················· |
| D | 7174 | hCoV-19/Belgium/rega-0423300/2020\|EPI_ISL_462154\|2 | ···················· | ···················· | ····················· | ···················· | ···················· | ···················· |
| D | 7175 | hCoV-19/Belgium/rega-0423301/2020\|EPI_ISL_462155\|2 | ···················· | ···················· | ····················· | ···················· | ···················· | ···················· |
| D | 7176 | hCoV-19/Belgium/rega-0423302/2020\|EPI_ISL_462156\|2 | ···················· | ···················· | ····················· | ···················· | ···················· | ···················· |
| D | 7177 | hCoV-19/Belgium/rega-0423303/2020\|EPI_ISL_462157\|2 | ···················· | ···················· | ····················· | ···················· | ···················· | ···················· |
| D | 7178 | hCoV-19/Belgium/RM-0330304/2020\|EPI_ISL_462158\|202 | ···················· | ···················· | ····················· | ···················· | ···················· | ···················· |
| D | 7179 | hCoV-19/Belgium/VDLM-0330305/2020\|EPI_ISL_462159\|2 | ···················· | ···················· | ····················· | ···················· | ···················· | ···················· |
| D | 7180 | hCoV-19/Belgium/DWW-0330306/2020\|EPI_ISL_462160\|20 | ···················· | ···················· | ····················· | ···················· | ···················· | ···················· |
| D | 7181 | hCoV-19/Belgium/DJS-0330307/2020\|EPI_ISL_462161\|20 | ···················· | ···················· | ····················· | ···················· | ···················· | ···················· |
| D | 7182 | hCoV-19/Belgium/VRS-0320308/2020\|EPI_ISL_462162\|20 | ···················· | ···················· | ····················· | ···················· | ···················· | ···················· |
| D | 7183 | hCoV-19/Belgium/GQL-0328309/2020\|EPI_ISL_462163\|20 | ···················· | ···················· | ····················· | ···················· | ···················· | ···················· |
| D | 7184 | hCoV-19/Belgium/LJ-0328310/2020\|EPI_ISL_462164\|202 | ···················· | ···················· | ····················· | ···················· | ···················· | ···················· |
| D | 7185 | hCoV-19/Belgium/FA-0328311/2020\|EPI_ISL_462165\|202 | ···················· | ···················· | ····················· | ···················· | ···················· | ···················· |
| D | 7186 | hCoV-19/Belgium/CFM-0328312/2020\|EPI_ISL_462166\|20 | ···················· | ···················· | ····················· | ···················· | ···················· | ···················· |
| D | 7187 | hCoV-19/Belgium/GC-0328313/2020\|EPI_ISL_462167\|202 | ···················· | ···················· | ····················· | ···················· | ···················· | ···················· |
| D | 7188 | hCoV-19/Belgium/SG-0328314/2020\|EPI_ISL_462168\|202 | ···················· | ···················· | ····················· | ···················· | ···················· | ···················· |
| D | 7189 | hCoV-19/Belgium/RR-0326315/2020\|EPI_ISL_462169\|202 | ···················· | ···················· | ····················· | ···················· | ···················· | ···················· |
| D | 7190 | hCoV-19/Belgium/PF-0326316/2020\|EPI_ISL_462170\|202 | ···················· | ·A·················· | ····················· | ···················· | ···················· | ···················· |
| D | 7191 | hCoV-19/Belgium/VJB-0327317/2020\|EPI_ISL_462171\|20 | ···················· | ···················· | ····················· | ···················· | ···················· | ···················· |
| D | 7192 | hCoV-19/Belgium/DAL-0327318/2020\|EPI_ISL_462172\|20 | ···················· | ···················· | ····················· | ···················· | ···················· | ···················· |
| D | 7193 | hCoV-19/Belgium/VA-0327319/2020\|EPI_ISL_462173\|202 | ···················· | ···················· | ····················· | ···················· | ···················· | ···················· |
| D | 7194 | hCoV-19/Belgium/BM-0327320/2020\|EPI_ISL_462174\|202 | ···················· | ···················· | ····················· | ···················· | ···················· | ···················· |
| D | 7195 | hCoV-19/Belgium/AF-0327321/2020\|EPI_ISL_462175\|202 | ···················· | ···················· | ····················· | ···················· | ···················· | ···················· |
| D | 7196 | hCoV-19/Belgium/IL-0326322/2020\|EPI_ISL_462176\|202 | ···················· | ···················· | ····················· | ···················· | ···················· | ···················· |
| D | 7197 | hCoV-19/Belgium/BA-0326323/2020\|EPI_ISL_462177\|202 | ···················· | ···················· | ····················· | ···················· | ···················· | ···················· |
| D | 7198 | hCoV-19/USA/WA-S910/2020\|EPI_ISL_463400\|2020-04-27 | ···················· | ···················· | ····················· | ···················· | ···················· | ···················· |
| D | 7199 | hCoV-19/USA/WA-S911/2020\|EPI_ISL_463401\|2020-04-28 | ···················· | ···················· | ····················· | ···················· | ···················· | ···················· |
| D | 7200 | hCoV-19/USA/WA-S912/2020\|EPI_ISL_463402\|2020-04-27 | ···················· | ···················· | ····················· | ···················· | ···················· | ···················· |
| D | 7201 | hCoV-19/USA/WA-S913/2020\|EPI_ISL_463403\|2020-04-27 | ···················· | ···················· | ····················· | ···················· | ···················· | ···················· |
| D | 7202 | hCoV-19/USA/WA-S916/2020\|EPI_ISL_463406\|2020-04-27 | ···················· | ···················· | ····················· | ···················· | ···················· | ···················· |
| D | 7203 | hCoV-19/USA/WA-S917/2020\|EPI_ISL_463407\|2020-04-27 | ···················· | ···················· | ····················· | ···················· | ···················· | ···················· |
| D | 7204 | hCoV-19/USA/WA-S918/2020\|EPI_ISL_463408\|2020-04-24 | ···················· | ···················· | ····················· | ···················· | ···················· | ···················· |
| D | 7205 | hCoV-19/USA/WA-S919/2020\|EPI_ISL_463409\|2020-04-25 | ···················· | ···················· | ····················· | ···················· | ···················· | ···················· |
| D | 7206 | hCoV-19/USA/WA-S920/2020\|EPI_ISL_463410\|2020-04-25 | ···················· | ···················· | ····················· | ···················· | ···················· | ···················· |
| D | 7207 | hCoV-19/USA/WA-S921/2020\|EPI_ISL_463411\|2020-04-25 | ···················· | ···················· | ····················· | ···················· | ···················· | ···················· |
| D | 7208 | hCoV-19/USA/WA-S922/2020\|EPI_ISL_463412\|2020-04-26 | ···················· | ···················· | ····················· | ···················· | ···················· | ···················· |
| D | 7209 | hCoV-19/USA/WA-S923/2020\|EPI_ISL_463413\|2020-04-26 | ···················· | ···················· | ····················· | ···················· | ···················· | ···················· |
| D | 7210 | hCoV-19/USA/WA-S924/2020\|EPI_ISL_463414\|2020-04-26 | ···················· | ···················· | ····················· | ···················· | ···················· | ···················· |
| D | 7211 | hCoV-19/USA/WA-S925/2020\|EPI_ISL_463415\|2020-04-24 | ···················· | ···················· | ····················· | ···················· | ···················· | ···················· |
| D | 7212 | hCoV-19/USA/WA-S926/2020\|EPI_ISL_463416\|2020-04-22 | ···················· | ···················· | ····················· | ···················· | ···················· | ···················· |
| D | 7213 | hCoV-19/USA/WA-S927/2020\|EPI_ISL_463417\|2020-04-26 | ···················· | ···················· | ····················· | ···················· | ···················· | ···················· |
| D | 7214 | hCoV-19/USA/WA-S928/2020\|EPI_ISL_463418\|2020-04-23 | ···················· | ···················· | ····················· | ···················· | ···················· | ···················· |
| D | 7215 | hCoV-19/USA/WA-S929/2020\|EPI_ISL_463419\|2020-04-23 | ···················· | ···················· | ····················· | ···················· | ···················· | ···················· |
| D | 7216 | hCoV-19/USA/WA-S930/2020\|EPI_ISL_463420\|2020-04-23 | ···················· | ···················· | ····················· | ···················· | ···················· | ···················· |
| D | 7217 | hCoV-19/USA/WA-S931/2020\|EPI_ISL_463421\|2020-04-23 | ···················· | ···················· | ····················· | ···················· | ···················· | ···················· |
| D | 7218 | hCoV-19/USA/WA-S932/2020\|EPI_ISL_463422\|2020-04-23 | ···················· | ···················· | ····················· | ···················· | ···················· | ···················· |
| D | 7219 | hCoV-19/USA/WA-S933/2020\|EPI_ISL_463423\|2020-04-23 | ···················· | ···················· | ····················· | ···················· | ···················· | ···················· |
| D | 7220 | hCoV-19/USA/WA-S934/2020\|EPI_ISL_463424\|2020-04-23 | ···················· | ···················· | ····················· | ···················· | ···················· | ···················· |
| D | 7221 | hCoV-19/USA/WA-S935/2020\|EPI_ISL_463425\|2020-04-23 | ···················· | ···················· | ····················· | ···················· | ···················· | ···················· |
| D | 7222 | hCoV-19/USA/WA-S936/2020\|EPI_ISL_463426\|2020-04-23 | ···················· | ···················· | ····················· | ···················· | ···················· | ···················· |
| D | 7223 | hCoV-19/USA/WA-S937/2020\|EPI_ISL_463427\|2020-04-23 | ···················· | ···················· | ····················· | ···················· | ···················· | ···················· |
| D | 7224 | hCoV-19/USA/WA-S938/2020\|EPI_ISL_463428\|2020-04-23 | ···················· | ···················· | ····················· | ···················· | ···················· | ···················· |
| D | 7225 | hCoV-19/USA/WA-S939/2020\|EPI_ISL_463429\|2020-04-23 | ···················· | ···················· | ····················· | ···················· | ···················· | ···················· |
| D | 7226 | hCoV-19/USA/WA-S940/2020\|EPI_ISL_463430\|2020-04-23 | ···················· | ···················· | ····················· | ···················· | ···················· | ···················· |
| D | 7227 | hCoV-19/USA/WA-S941/2020\|EPI_ISL_463431\|2020-04-23 | ···················· | ···················· | ····················· | ···················· | ···················· | ···················· |
| D | 7228 | hCoV-19/USA/WA-S942/2020\|EPI_ISL_463432\|2020-04-27 | ···················· | ···················· | ····················· | ···················· | ···················· | ···················· |

| | | | | | | | |
|---|---|---|---|---|---|---|---|
| D | 7229 hCoV-19/USA/WA-S943/2020\|EPI_ISL_463433\|2020-04-27 | ···················· | ···················· | ···················· | ···················· | ···················· | ···················· |
| D | 7230 hCoV-19/USA/WA-S944/2020\|EPI_ISL_463434\|2020-04-27 | ···················· | ···················· | ···················· | ···················· | ···················· | ···················· |
| D | 7231 hCoV-19/USA/WA-S945/2020\|EPI_ISL_463435\|2020-04-27 | ···················· | ···················· | ···················· | ···················· | ···················· | ···················· |
| D | 7232 hCoV-19/USA/WA-S946/2020\|EPI_ISL_463436\|2020-04-27 | ···················· | ···················· | ···················· | ···················· | ···················· | ···················· |
| D | 7233 hCoV-19/USA/WA-S947/2020\|EPI_ISL_463437\|2020-04-26 | ···················· | ···················· | ···················· | ···················· | ···················· | ···················· |
| D | 7234 hCoV-19/USA/WA-S948/2020\|EPI_ISL_463438\|2020-04-26 | ···················· | ···················· | ···················· | ···················· | ···················· | ···················· |
| D | 7235 hCoV-19/USA/WA-S949/2020\|EPI_ISL_463439\|2020-04-27 | ···················· | ···················· | ···················· | ···················· | ···················· | ···················· |
| D | 7236 hCoV-19/USA/WA-S950/2020\|EPI_ISL_463440\|2020-04-27 | ···················· | ···················· | ···················· | ···················· | ···················· | ···················· |
| D | 7237 hCoV-19/USA/WA-S951/2020\|EPI_ISL_463441\|2020-04-27 | ···················· | ···················· | ···················· | ···················· | ···················· | ···················· |
| D | 7238 hCoV-19/USA/WA-S952/2020\|EPI_ISL_463442\|2020-04-27 | ···················· | ···················· | ···················· | ···················· | ···················· | ···················· |
| D | 7239 hCoV-19/USA/WA-S988/2020\|EPI_ISL_463478\|2020-04-28 | ···················· | ···················· | ···················· | ···················· | ···················· | ···················· |
| D | 7240 hCoV-19/USA/WA-S989/2020\|EPI_ISL_463479\|2020-04-28 | ···················· | ···················· | ···················· | ···················· | ···················· | ···················· |
| D | 7241 hCoV-19/USA/WA-S990/2020\|EPI_ISL_463480\|2020-04-28 | ···················· | ···················· | ···················· | ···················· | ···················· | ···················· |
| D | 7242 hCoV-19/USA/WA-S991/2020\|EPI_ISL_463481\|2020-04-27 | ···················· | ···················· | ···················· | ···················· | ···················· | ···················· |
| D | 7243 hCoV-19/USA/WA-S992/2020\|EPI_ISL_463482\|2020-04-28 | ···················· | ···················· | ···················· | ···················· | ···················· | ···················· |
| D | 7244 hCoV-19/USA/WA-S993/2020\|EPI_ISL_463483\|2020-04-29 | ···················· | ···················· | ···················· | ···················· | ···················· | ···················· |
| D | 7245 hCoV-19/USA/WA-S994/2020\|EPI_ISL_463484\|2020-04-29 | ···················· | ···················· | ···················· | ···················· | ···················· | ···················· |
| D | 7246 hCoV-19/USA/WA-S995/2020\|EPI_ISL_463485\|2020-04-29 | ···················· | ···················· | ···················· | ···················· | ···················· | ···················· |
| D | 7247 hCoV-19/USA/WA-S996/2020\|EPI_ISL_463486\|2020-04-28 | ···················· | ···················· | ···················· | ···················· | ···················· | ···················· |
| D | 7248 hCoV-19/USA/WA-S997/2020\|EPI_ISL_463487\|2020-04-28 | ···················· | ···················· | ···················· | ···················· | ···················· | ···················· |
| D | 7249 hCoV-19/USA/WA-S998/2020\|EPI_ISL_463488\|2020-04-28 | ···················· | ···················· | ···················· | ···················· | ···················· | ···················· |
| D | 7250 hCoV-19/USA/WA-S999/2020\|EPI_ISL_463489\|2020-04-28 | ···················· | ···················· | ···················· | ···················· | ···················· | ···················· |
| D | 7251 hCoV-19/USA/WA-S1000/2020\|EPI_ISL_463490\|2020-04-2 | ···················· | ···················· | ···················· | ···················· | ···················· | ···················· |
| D | 7252 hCoV-19/USA/WA-S1001/2020\|EPI_ISL_463491\|2020-04-2 | ···················· | ···················· | ···················· | ···················· | ···················· | ···················· |
| D | 7253 hCoV-19/USA/WA-S1002/2020\|EPI_ISL_463492\|2020-04-2 | ···················· | ···················· | ···················· | ···················· | ···················· | ···················· |
| D | 7254 hCoV-19/USA/WA-S1003/2020\|EPI_ISL_463493\|2020-04-2 | ···················· | ···················· | ···················· | ···················· | ···················· | ···················· |
| D | 7255 hCoV-19/USA/WA-S1004/2020\|EPI_ISL_463494\|2020-04-2 | ···················· | ···················· | ···················· | ···················· | ···················· | ···················· |
| D | 7256 hCoV-19/USA/WA-S1005/2020\|EPI_ISL_463495\|2020-04-2 | ···················· | ···················· | ···················· | ···················· | ···················· | ···················· |
| D | 7257 hCoV-19/USA/WA-S1006/2020\|EPI_ISL_463496\|2020-04-2 | ···················· | ···················· | ···················· | ···················· | ···················· | ···················· |
| D | 7258 hCoV-19/USA/WA-S1007/2020\|EPI_ISL_463497\|2020-04-3 | ···················· | ···················· | ···················· | ···················· | ···················· | ···················· |
| D | 7259 hCoV-19/USA/WA-S1008/2020\|EPI_ISL_463498\|2020-04-2 | ···················· | ···················· | ···················· | ···················· | ···················· | ···················· |
| D | 7260 hCoV-19/USA/WA-S1009/2020\|EPI_ISL_463499\|2020-04-3 | ···················· | ···················· | ···················· | ···················· | ···················· | ···················· |
| D | 7261 hCoV-19/England/201140388/2020\|EPI_ISL_464730\|2020 | ···················· | ···················· | ···················· | ···················· | ···················· | ···················· |
| D | 7263 hCoV-19/England/201140432/2020\|EPI_ISL_464734\|2020 | ···················· | ···················· | ···················· | ···················· | ···················· | ···················· |
| D | 7267 hCoV-19/England/201141047/2020\|EPI_ISL_464742\|2020 | ···················· | ···················· | ···················· | ···················· | ···················· | ···················· |
| D | 7268 hCoV-19/England/201141063/2020\|EPI_ISL_464743\|2020 | ···················· | ···················· | ···················· | ···················· | ···················· | ···················· |
| D | 7270 hCoV-19/England/201141179/2020\|EPI_ISL_464745\|2020 | ···················· | ···················· | ···················· | ···················· | ···················· | ···················· |
| D | 7272 hCoV-19/England/201141471/2020\|EPI_ISL_464755\|2020 | ···················· | ···················· | ···················· | ···················· | ···················· | ···················· |
| D | 7274 hCoV-19/England/201141520/2020\|EPI_ISL_464758\|2020 | ···················· | ···················· | ···················· | ···················· | ···················· | ·············NN····· |
| D | 7277 hCoV-19/England/201141550/2020\|EPI_ISL_464765\|2020 | ···················· | ···················· | ···················· | ···················· | ···················· | ··NN················ |
| D | 7278 hCoV-19/England/201141596/2020\|EPI_ISL_464767\|2020 | ···················· | ···················· | ···················· | ···················· | ···················· | ···················· |
| D | 7280 hCoV-19/England/201160002/2020\|EPI_ISL_464770\|2020 | ···················· | ···················· | ···················· | ···················· | ···················· | ···················· |
| D | 7283 hCoV-19/England/BRIS-12847E/2020\|EPI_ISL_469600\|20 | ···················· | ···················· | ···················· | ···················· | ···················· | ···················· |
| D | 7284 hCoV-19/England/BRIS-1276A6/2020\|EPI_ISL_469603\|20 | ···················· | ···················· | ···················· | ···················· | ···················· | ···················· |
| D | 7285 hCoV-19/England/BRIS-12846F/2020\|EPI_ISL_469604\|20 | ···················· | ···················· | ···················· | ···················· | ···················· | ···················· |
| D | 7286 hCoV-19/England/BRIS-127891/2020\|EPI_ISL_469605\|20 | ···················· | ···················· | ···················· | ···················· | ···················· | ···················· |
| D | 7287 hCoV-19/England/BRIS-127FE7/2020\|EPI_ISL_469607\|20 | ···················· | ·A·················· | ···················· | ···················· | ···················· | ···················· |
| D | 7288 hCoV-19/England/BRIS-12799E/2020\|EPI_ISL_469608\|20 | ···················· | ···················· | ···················· | ···················· | ···················· | ···················· |
| D | 7289 hCoV-19/England/BRIS-127873/2020\|EPI_ISL_469609\|20 | ···················· | ···················· | ···················· | ···················· | ···················· | ···················· |
| D | 7290 hCoV-19/England/BRIS-127275/2020\|EPI_ISL_469610\|20 | ···················· | ·············A······ | ···················· | ···················· | ···················· | ···················· |
| D | 7291 hCoV-19/England/BRIS-1282CF/2020\|EPI_ISL_469612\|20 | ···················· | ···················· | ···················· | ···················· | ···················· | ···················· |
| D | 7292 hCoV-19/England/BRIS-127FAB/2020\|EPI_ISL_469613\|20 | ···················· | ····A··············· | ···················· | ···················· | ···················· | ···················· |
| D | 7293 hCoV-19/England/BRIS-127239/2020\|EPI_ISL_469614\|20 | ···················· | ···················· | ···················· | ···················· | ···················· | ···················· |
| D | 7294 hCoV-19/England/BRIS-128274/2020\|EPI_ISL_469615\|20 | ···················· | ···················· | ···················· | ···················· | ···················· | ···················· |
| D | 7295 hCoV-19/England/BRIS-128432/2020\|EPI_ISL_469616\|20 | ···················· | ···················· | ···················· | ···················· | ···················· | ···················· |
| D | 7296 hCoV-19/England/BRIS-1280A7/2020\|EPI_ISL_469621\|20 | ···················· | ···················· | ···················· | ···················· | ···················· | ···················· |
| D | 7297 hCoV-19/England/BRIS-127372/2020\|EPI_ISL_469624\|20 | ···················· | ···················· | ···················· | ···················· | ···················· | ···················· |
| D | 7298 hCoV-19/England/EXET-136D72/2020\|EPI_ISL_471921\|20 | ···················· | ···················· | ···················· | ···················· | ···················· | ···················· |
| D | 7299 hCoV-19/England/EXET-136D81/2020\|EPI_ISL_471922\|20 | ···················· | ···················· | ···················· | ···················· | ···················· | ···················· |
| D | 7300 hCoV-19/England/EXET-136DAF/2020\|EPI_ISL_471924\|20 | ···················· | ···················· | ···················· | ···················· | ···················· | ···················· |
| D | 7301 hCoV-19/England/EXET-136DDC/2020\|EPI_ISL_471925\|20 | ···················· | ···················· | ···················· | ···················· | ···················· | ···················· |
| D | 7302 hCoV-19/England/EXET-136DEB/2020\|EPI_ISL_471926\|20 | ···················· | ···················· | ···················· | ···················· | ···················· | ···················· |
| D | 7303 hCoV-19/England/EXET-136E33/2020\|EPI_ISL_471927\|20 | ···················· | ···················· | ···················· | ···················· | ···················· | ···················· |
| D | 7304 hCoV-19/England/EXET-136E9D/2020\|EPI_ISL_471929\|20 | ···················· | ···················· | ···················· | ···················· | ···················· | ···················· |
| D | 7305 hCoV-19/England/EXET-136EAC/2020\|EPI_ISL_471930\|20 | ···················· | ···················· | ···················· | ···················· | ···················· | ···················· |
| D | 7306 hCoV-19/England/EXET-136EBB/2020\|EPI_ISL_471931\|20 | ···················· | ···················· | ···················· | ···················· | ···················· | ···················· |
| D | 7307 hCoV-19/England/EXET-136ECA/2020\|EPI_ISL_471932\|20 | ···················· | ···················· | ···················· | ···················· | ···················· | ···················· |
| D | 7308 hCoV-19/England/EXET-136ED9/2020\|EPI_ISL_471933\|20 | ···················· | ···················· | ···················· | ···················· | ···················· | ···················· |
| D | 7309 hCoV-19/England/EXET-136EE8/2020\|EPI_ISL_471934\|20 | ···················· | ···················· | ···················· | ···················· | ···················· | ···················· |
| D | 7310 hCoV-19/England/EXET-136F30/2020\|EPI_ISL_471935\|20 | ···················· | ···················· | ···················· | ···················· | ···················· | ···················· |
| D | 7311 hCoV-19/England/EXET-136F4F/2020\|EPI_ISL_471936\|20 | ···················· | ···················· | ···················· | ···················· | ···················· | ···················· |
| D | 7312 hCoV-19/England/EXET-136F5E/2020\|EPI_ISL_471937\|20 | ···················· | ···················· | ···················· | ···················· | ···················· | ···················· |
| D | 7313 hCoV-19/England/EXET-136F6D/2020\|EPI_ISL_471938\|20 | ···················· | ···················· | ···················· | ···················· | ···················· | ···················· |
| D | 7314 hCoV-19/Wales/PHWC-163EE2/2020\|EPI_ISL_474100\|2020 | ···················· | ···················· | ···················· | ···················· | ···················· | ···················· |
| D | 7315 hCoV-19/Wales/PHWC-163EF1/2020\|EPI_ISL_474101\|2020 | ···················· | ···················· | ···················· | ···················· | ···················· | ···················· |
| D | 7316 hCoV-19/Wales/PHWC-163F2B/2020\|EPI_ISL_474103\|2020 | ···················· | ···················· | ···················· | ···················· | ···················· | ···················· |
| D | 7317 hCoV-19/Wales/PHWC-163F3A/2020\|EPI_ISL_474104\|2020 | ···················· | ···················· | ···················· | ···················· | ···················· | ···················· |
| D | 7318 hCoV-19/Wales/PHWC-163F49/2020\|EPI_ISL_474105\|2020 | ···················· | ···················· | ···················· | ···················· | ···················· | ···················· |

D 7319 hCoV-19/Wales/PHWC-163F94/2020|EPI_ISL_474110|2020 ···················· ···················· ····················· ···················· ···················· ····················
D 7320 hCoV-19/Wales/PHWC-163FB2/2020|EPI_ISL_474112|2020 ···················· ···················· ····················· ···················· ···················· ····················
D 7321 hCoV-19/Wales/PHWC-163FC1/2020|EPI_ISL_474113|2020 ···················· ···················· ····················· ···················· ···················· ····················
D 7322 hCoV-19/Wales/PHWC-164009/2020|EPI_ISL_474115|2020 ···················· ···················· ····················· ···················· ···················· ····················
D 7323 hCoV-19/Wales/PHWC-164018/2020|EPI_ISL_474116|2020 ···················· ···················· ····················· ···················· ···················· ····················
D 7324 hCoV-19/Wales/PHWC-164027/2020|EPI_ISL_474117|2020 ···················· ···················· ····················· ···················· ···················· ····················
D 7325 hCoV-19/Wales/PHWC-164036/2020|EPI_ISL_474118|2020 ···················· ···················· ····················· ···················· ···················· ····················
D 7326 hCoV-19/Wales/PHWC-164072/2020|EPI_ISL_474122|2020 ···················· ···················· ····················· ···················· ···················· ····················
D 7327 hCoV-19/Wales/PHWC-164090/2020|EPI_ISL_474124|2020 ···················· ···················· ····················· ···················· ···················· ····················
D 7328 hCoV-19/Wales/PHWC-1640AF/2020|EPI_ISL_474125|2020 ···················· ···················· ····················· ···················· ···················· ····················
D 7329 hCoV-19/Wales/PHWC-1640BE/2020|EPI_ISL_474126|2020 ···················· ···················· ····················· ···················· ···················· ····················
D 7330 hCoV-19/Wales/PHWC-164124/2020|EPI_ISL_474131|2020 ···················· ···················· ····················· ···················· ···················· ····················
D 7332 hCoV-19/Wales/PHWC-16417F/2020|EPI_ISL_474136|2020 ···················· ···················· ····················· ···················· ···················· ····················
D 7333 hCoV-19/Wales/PHWC-16418E/2020|EPI_ISL_474137|2020 ···················· ···················· ····················· ···················· ···················· ····················
D 7334 hCoV-19/Wales/PHWC-1641BB/2020|EPI_ISL_474140|2020 ···················· ···················· ····················· ···················· ···················· ····················
D 7335 hCoV-19/Wales/PHWC-1641CA/2020|EPI_ISL_474141|2020 ···················· ···················· ····················· ···················· ···················· ····················
D 7336 hCoV-19/Wales/PHWC-164230/2020|EPI_ISL_474147|2020 ···················· ···················· ····················· ···················· ···················· ····················
D 7337 hCoV-19/Wales/PHWC-16426D/2020|EPI_ISL_474150|2020 ···················· ···················· ····················· ···················· ···················· ····················
D 7338 hCoV-19/Wales/PHWC-16428B/2020|EPI_ISL_474152|2020 ···················· ···················· ····················· ···················· ···················· ····················
D 7339 hCoV-19/Wales/PHWC-1642C7/2020|EPI_ISL_474155|2020 ···················· ···················· ····················· ···················· ···················· ····················
D 7340 hCoV-19/Wales/PHWC-1642D6/2020|EPI_ISL_474156|2020 ···················· ···················· ····················· ···················· ···················· ····················
D 7341 hCoV-19/Wales/PHWC-1642E5/2020|EPI_ISL_474157|2020 ···················· ···················· ····················· ···················· ···················· ····················
D 7342 hCoV-19/Wales/PHWC-164300/2020|EPI_ISL_474159|2020 ···················· ···················· ····················· ···················· ···················· ····················
D 7343 hCoV-19/Wales/PHWC-16431F/2020|EPI_ISL_474160|2020 ···················· ···················· ····················· ···················· ···················· ····················
D 7344 hCoV-19/Wales/PHWC-16432E/2020|EPI_ISL_474161|2020 ···················· ···················· ····················· ···················· ···················· ····················
D 7345 hCoV-19/Wales/PHWC-16434C/2020|EPI_ISL_474162|2020 ···················· ···················· ····················· ···················· ···················· ····················
D 7346 hCoV-19/Wales/PHWC-16436A/2020|EPI_ISL_474164|2020 ···················· ···················· ····················· ···················· ···················· ····················
D 7347 hCoV-19/Wales/PHWC-164379/2020|EPI_ISL_474165|2020 ···················· ···················· ····················· ···················· ···················· ····················
D 7348 hCoV-19/Wales/PHWC-164397/2020|EPI_ISL_474167|2020 ···················· ···················· ····················· ···················· ···················· ····················
D 7349 hCoV-19/Wales/PHWC-1643A6/2020|EPI_ISL_474168|2020 ···················· ···················· ····················· ···················· ···················· ····················
D 7350 hCoV-19/Wales/PHWC-1643B5/2020|EPI_ISL_474169|2020 ···················· ···················· ····················· ···················· ···················· ····················
D 7351 hCoV-19/Wales/PHWC-1643D3/2020|EPI_ISL_474171|2020 ···················· ···················· ····················· ···················· ···················· ····················
D 7352 hCoV-19/Wales/PHWC-1643F1/2020|EPI_ISL_474173|2020 ···················· ···················· ····················· ···················· ···················· ····················
D 7353 hCoV-19/Wales/PHWC-16441C/2020|EPI_ISL_474175|2020 ···················· ···················· ····················· ···················· ···················· ····················
D 7354 hCoV-19/Wales/PHWC-16443A/2020|EPI_ISL_474177|2020 ···················· ···················· ····················· ···················· ···················· ····················
D 7355 hCoV-19/Wales/PHWC-164449/2020|EPI_ISL_474178|2020 ···················· ···················· ····················· ···················· ···················· ····················
D 7356 hCoV-19/Wales/PHWC-164555/2020|EPI_ISL_474193|2020 ···················· ···················· ····················· ···················· ···················· ····················
D 7357 hCoV-19/Wales/PHWC-164573/2020|EPI_ISL_474195|2020 ···················· ···················· ····················· ···················· ···················· ····················
D 7358 hCoV-19/England/SHEF-C9962/2020|EPI_ISL_475423|202 ···················· ···················· ····················· ···················· ···················· ····················
D 7359 hCoV-19/England/SHEF-C999F/2020|EPI_ISL_475424|202 ···················· ···················· ····················· ···················· ···················· ····················
D 7360 hCoV-19/England/SHEF-C9A23/2020|EPI_ISL_475425|202 ···················· ···················· ····················· ···················· ···················· ····················
D 7361 hCoV-19/England/SHEF-CA187/2020|EPI_ISL_475429|202 ···················· ···················· ····················· ···················· ···················· ····················
D 7362 hCoV-19/England/SHEF-CB0D4/2020|EPI_ISL_475434|202 ···················· ···················· ····················· ···················· ···················· ····················
D 7363 hCoV-19/England/SHEF-CC1C1/2020|EPI_ISL_475443|202 ···················· ···················· ····················· ···················· ···················· ····················
D 7364 hCoV-19/England/SHEF-CC440/2020|EPI_ISL_475445|202 ···················· ···················· ····················· ···················· ···················· ····················
D 7365 hCoV-19/England/SHEF-CCAC6/2020|EPI_ISL_475447|202 ···················· ···················· ····················· ···················· ···················· ····················
D 7366 hCoV-19/England/SHEF-CCCEE/2020|EPI_ISL_475448|202 ···················· ···················· ····················· ···················· ···················· ····················
D 7367 hCoV-19/England/SHEF-CD1EE/2020|EPI_ISL_475453|202 ···················· ···················· ····················· ···················· ···················· ····················
D 7368 hCoV-19/England/SHEF-CD245/2020|EPI_ISL_475454|202 ···················· ···················· ····················· ···················· ···················· ····················
D 7369 hCoV-19/England/SHEF-CD4B8/2020|EPI_ISL_475456|202 ···················· ···················· ····················· ···················· ···················· ····················
D 7370 hCoV-19/England/SHEF-CD588/2020|EPI_ISL_475457|202 ···················· ···················· ····················· ···················· ···················· ····················
D 7371 hCoV-19/England/SHEF-CE00E/2020|EPI_ISL_475461|202 ···················· ···················· ····················· ···················· ···················· ···············A····
D 7372 hCoV-19/England/SHEF-CE7FA/2020|EPI_ISL_475466|202 ···················· ···················· ····················· ···················· ···················· ····················
D 7373 hCoV-19/England/SHEF-CE8D9/2020|EPI_ISL_475469|202 ···················· ···················· ····················· ···················· ···················· ····················
D 7374 hCoV-19/England/SHEF-CEC37/2020|EPI_ISL_475471|202 ···················· ···················· ····················· ···················· ···················· ····················
D 7375 hCoV-19/England/SHEF-CED25/2020|EPI_ISL_475473|202 ···················· ···················· ····················· ···················· ···················· ····················
D 7376 hCoV-19/England/SHEF-CF270/2020|EPI_ISL_475476|202 ···················· ···················· ····················· ···················· ···················· ····················
D 7377 hCoV-19/England/SHEF-CF498/2020|EPI_ISL_475477|202 ···················· ···················· ····················· ···················· ···················· ····················
D 7378 hCoV-19/England/SHEF-CF4F2/2020|EPI_ISL_475478|202 ···················· ···················· ····················· ···················· ···················· ····················
D 7379 hCoV-19/England/SHEF-CFD42/2020|EPI_ISL_475484|202 ···················· ···················· ····················· ···················· ···················· ····················
D 7380 hCoV-19/England/SHEF-CFE5E/2020|EPI_ISL_475485|202 ···················· ···················· ····················· ···················· ···················· ····················
D 7381 hCoV-19/England/SHEF-D0DA8/2020|EPI_ISL_475492|202 ···················· ···················· ····················· ···················· ···················· ····················
D 7382 hCoV-19/England/SHEF-D1976/2020|EPI_ISL_475497|202 ···················· ···················· ····················· ···················· ···················· ····················
D 7383 hCoV-19/USA/MN-MDH-1125/2020|EPI_ISL_476710|2020-0 ···················· ···················· ····················· ···················· ···················· ····················
D 7384 hCoV-19/USA/MN-MDH-1126/2020|EPI_ISL_476711|2020-0 ···················· ···················· ····················· ···················· ···················· ····················
D 7385 hCoV-19/USA/MN-MDH-1127/2020|EPI_ISL_476712|2020-0 ···················· ···················· ····················· ···················· ···················· ····················
D 7386 hCoV-19/USA/MN-MDH-1128/2020|EPI_ISL_476713|2020-0 ···················· ···················· ····················· ···················· ···················· ····················
D 7387 hCoV-19/USA/MN-MDH-1129/2020|EPI_ISL_476714|2020-0 ···················· ···················· ····················· ···················· ···················· ····················
D 7388 hCoV-19/USA/MN-MDH-1130/2020|EPI_ISL_476715|2020-0 ···················· ···················· ····················· ···················· ···················· ····················
D 7389 hCoV-19/USA/MN-MDH-1131/2020|EPI_ISL_476716|2020-0 ···················· ···················· ····················· ···················· ···················· ····················
D 7390 hCoV-19/USA/MN-MDH-1132/2020|EPI_ISL_476717|2020-0 ···················· ···················· ····················· ···················· ···················· ····················
D 7391 hCoV-19/USA/MN-MDH-1133/2020|EPI_ISL_476718|2020-0 ···················· ···················· ····················· ···················· ···················· ····················
D 7392 hCoV-19/USA/MN-MDH-1134/2020|EPI_ISL_476719|2020-0 ···················· ···················· ····················· ···················· ···················· ····················
D 7393 hCoV-19/USA/MN-MDH-1141/2020|EPI_ISL_476726|2020-0 ···················· ···················· ····················· ···················· ···················· ····················
D 7394 hCoV-19/USA/MN-MDH-1168/2020|EPI_ISL_476753|2020-0 ···················· ···················· ····················· ···················· ···················· ····················
D 7395 hCoV-19/USA/MN-MDH-1169/2020|EPI_ISL_476754|2020-0 ···················· ···················· ····················· ···················· ···················· ····················
D 7396 hCoV-19/USA/MN-MDH-1170/2020|EPI_ISL_476755|2020-0 ···················· ···················· ····················· ···················· ···················· ····················
D 7397 hCoV-19/USA/MN-MDH-1171/2020|EPI_ISL_476756|2020-0 ···················· ···················· ····················· ···················· ···················· ····················

| | | | | | | | | |
|---|---|---|---|---|---|---|---|---|
| D | 7398 hCoV-19/USA/MN-MDH-1172/2020\|EPI_ISL_476757\|2020-0 | ···················· | ·A·················· | ···················· | ···················· | ···················· | ···················· | |
| D | 7399 hCoV-19/USA/MN-MDH-1173/2020\|EPI_ISL_476758\|2020-0 | ···················· | ·A·················· | ···················· | ···················· | ···················· | ···················· | |
| D | 7400 hCoV-19/USA/MN-MDH-1174/2020\|EPI_ISL_476759\|2020-0 | ···················· | ·A·················· | ···················· | ···················· | ···················· | ···················· | |
| D | 7401 hCoV-19/USA/MN-MDH-1175/2020\|EPI_ISL_476760\|2020-0 | ···················· | ·A·················· | ···················· | ···················· | ···················· | ···················· | |
| D | 7402 hCoV-19/USA/MN-MDH-1176/2020\|EPI_ISL_476761\|2020-0 | ···················· | ·A·················· | ···················· | ···················· | ···················· | ···················· | |
| D | 7403 hCoV-19/USA/MN-MDH-1177/2020\|EPI_ISL_476762\|2020-0 | ···················· | ···················· | ···················· | ···················· | ···················· | ···················· | |
| D | 7404 hCoV-19/USA/MN-MDH-1178/2020\|EPI_ISL_476763\|2020-0 | ···················· | ···················· | ···················· | ···················· | ···················· | ···················· | |
| D | 7405 hCoV-19/USA/MN-MDH-1179/2020\|EPI_ISL_476764\|2020-0 | ···················· | ···················· | ···················· | ···················· | ···················· | ···················· | |
| D | 7406 hCoV-19/USA/MN-MDH-1180/2020\|EPI_ISL_476765\|2020-0 | ···················· | ···················· | ···················· | ···················· | ···················· | ···················· | |
| D | 7407 hCoV-19/USA/MN-MDH-1181/2020\|EPI_ISL_476766\|2020-0 | ···················· | ···················· | ···················· | ···················· | ···················· | ···················· | |
| D | 7408 hCoV-19/USA/CA-CZB-1751/2020\|EPI_ISL_476768\|2020-0 | ···················· | ···················· | ···················· | ···················· | ···················· | ···················· | |
| D | 7409 hCoV-19/USA/CA-CZB-1755/2020\|EPI_ISL_476772\|2020-0 | ···················· | ···················· | ···················· | ···················· | ···················· | ···················· | |
| D | 7410 hCoV-19/USA/CA-CZB-1756/2020\|EPI_ISL_476773\|2020-0 | ···················· | ···················· | ···················· | ···················· | ···················· | ···················· | |
| D | 7411 hCoV-19/USA/CA-CZB-1757/2020\|EPI_ISL_476774\|2020-0 | ···················· | ···················· | ···················· | ···················· | ···················· | ···················· | |
| D | 7412 hCoV-19/USA/CA-CZB-1758/2020\|EPI_ISL_476775\|2020-0 | ···················· | ···················· | ···················· | ···················· | ···················· | ···················· | |
| D | 7413 hCoV-19/USA/CA-CZB-1761/2020\|EPI_ISL_476778\|2020-0 | ···················· | ···················· | ···················· | ···················· | ···················· | ···················· | |
| D | 7414 hCoV-19/USA/CA-CZB-1763/2020\|EPI_ISL_476780\|2020-0 | ···················· | ···················· | ···················· | ···················· | ···················· | ···················· | |
| D | 7415 hCoV-19/USA/CA-CZB-1764/2020\|EPI_ISL_476781\|2020-0 | ···················· | ···················· | ···················· | ···················· | ···················· | ···················· | |
| D | 7416 hCoV-19/USA/CA-CZB-1765/2020\|EPI_ISL_476782\|2020-0 | ···················· | ···················· | ···················· | ···················· | ···················· | ···················· | |
| D | 7417 hCoV-19/USA/CA-CZB-1766/2020\|EPI_ISL_476783\|2020-0 | ···················· | ···················· | ···················· | ···················· | ···················· | ···················· | |
| D | 7418 hCoV-19/USA/CA-CZB-1767/2020\|EPI_ISL_476784\|2020-0 | ···················· | ···················· | ···················· | ···················· | ···················· | ···················· | |
| D | 7419 hCoV-19/USA/CA-CZB-1768/2020\|EPI_ISL_476785\|2020-0 | ···················· | ···················· | ···················· | ···················· | ···················· | ···················· | |
| D | 7420 hCoV-19/USA/CA-CZB-1769/2020\|EPI_ISL_476786\|2020-0 | ···················· | ···················· | ···················· | ···················· | ···················· | ···················· | |
| D | 7421 hCoV-19/USA/CA-CZB-1770/2020\|EPI_ISL_476787\|2020-0 | ···················· | ···················· | ···················· | ···················· | ···················· | ···················· | |
| D | 7422 hCoV-19/USA/CA-CZB-1771/2020\|EPI_ISL_476788\|2020-0 | ···················· | ···················· | ···················· | ···················· | ···················· | ···················· | |
| D | 7423 hCoV-19/USA/CA-CZB-1774/2020\|EPI_ISL_476791\|2020-0 | ···················· | ···················· | ···················· | ···················· | ···················· | ···················· | |
| D | 7424 hCoV-19/USA/CA-CZB-1775/2020\|EPI_ISL_476792\|2020-0 | ···················· | ···················· | ···················· | ···················· | ···················· | ···················· | |
| D | 7425 hCoV-19/USA/WA-S953/2020\|EPI_ISL_463443\|2020-04-27 | ···················· | ···················· | ···················· | ···················· | ···················· | ···················· | |
| D | 7426 hCoV-19/USA/WA-S954/2020\|EPI_ISL_463444\|2020-05-01 | ···················· | ···················· | ···················· | ···················· | ···················· | ···················· | |
| D | 7427 hCoV-19/USA/WA-S956/2020\|EPI_ISL_463446\|2020-05-01 | ···················· | ···················· | ···················· | ···················· | ···················· | ···················· | |
| D | 7428 hCoV-19/USA/WA-S959/2020\|EPI_ISL_463449\|2020-05-01 | ···················· | ···················· | ···················· | ···················· | ···················· | ···················· | |
| D | 7429 hCoV-19/USA/WA-S960/2020\|EPI_ISL_463450\|2020-05-01 | ···················· | ···················· | ···················· | ···················· | ···················· | ···················· | |
| D | 7430 hCoV-19/USA/WA-S961/2020\|EPI_ISL_463451\|2020-05-01 | ···················· | ···················· | ···················· | ···················· | ···················· | ···················· | |
| D | 7431 hCoV-19/USA/WA-S962/2020\|EPI_ISL_463452\|2020-05-01 | ···················· | ···················· | ···················· | ···················· | ···················· | ···················· | |
| D | 7432 hCoV-19/USA/WA-S963/2020\|EPI_ISL_463453\|2020-05-01 | ···················· | ···················· | ···················· | ···················· | ···················· | ···················· | |
| D | 7433 hCoV-19/USA/WA-S964/2020\|EPI_ISL_463454\|2020-05-01 | ···················· | ···················· | ···················· | ···················· | ···················· | ···················· | |
| D | 7434 hCoV-19/USA/WA-S965/2020\|EPI_ISL_463455\|2020-05-01 | ···················· | ···················· | ···················· | ···················· | ···················· | ···················· | |
| D | 7435 hCoV-19/USA/WA-S966/2020\|EPI_ISL_463456\|2020-04-30 | ···················· | ···················· | ···················· | ···················· | ···················· | ···················· | |
| D | 7436 hCoV-19/USA/WA-S967/2020\|EPI_ISL_463457\|2020-05-01 | ···················· | ···················· | ···················· | ···················· | ···················· | ···················· | |
| D | 7437 hCoV-19/USA/WA-S968/2020\|EPI_ISL_463458\|2020-05-01 | ···················· | ···················· | ···················· | ···················· | ···················· | ···················· | |
| D | 7438 hCoV-19/USA/WA-S969/2020\|EPI_ISL_463459\|2020-05-01 | ···················· | ···················· | ···················· | ···················· | ···················· | ···················· | |
| D | 7439 hCoV-19/USA/WA-S970/2020\|EPI_ISL_463460\|2020-05-01 | ···················· | ···················· | ···················· | ···················· | ···················· | ···················· | |
| D | 7440 hCoV-19/USA/WA-S971/2020\|EPI_ISL_463461\|2020-05-01 | ···················· | ···················· | ···················· | ···················· | ···················· | ···················· | |
| D | 7441 hCoV-19/USA/WA-S972/2020\|EPI_ISL_463462\|2020-05-01 | ···················· | ···················· | ···················· | ···················· | ···················· | ···················· | |
| D | 7442 hCoV-19/USA/WA-S973/2020\|EPI_ISL_463463\|2020-05-01 | ···················· | ···················· | ···················· | ···················· | ···················· | ···················· | |
| D | 7443 hCoV-19/USA/WA-S974/2020\|EPI_ISL_463464\|2020-05-01 | ···················· | ···················· | ···················· | ···················· | ···················· | ···················· | |
| D | 7444 hCoV-19/USA/WA-S975/2020\|EPI_ISL_463465\|2020-05-01 | ···················· | ···················· | ···················· | ···················· | ···················· | ···················· | |
| D | 7445 hCoV-19/USA/WA-S976/2020\|EPI_ISL_463466\|2020-04-28 | ···················· | ···················· | ···················· | ···················· | ···················· | ···················· | |
| D | 7446 hCoV-19/USA/WA-S977/2020\|EPI_ISL_463467\|2020-05-01 | ···················· | ···················· | ···················· | ···················· | ···················· | ···················· | |
| D | 7447 hCoV-19/USA/WA-S982/2020\|EPI_ISL_463472\|2020-04-28 | ···················· | ···················· | ···················· | ···················· | ···················· | ···················· | |
| D | 7448 hCoV-19/England/201140059/2020\|EPI_ISL_464711\|2020 | ···················· | ···················· | ···················· | ···················· | ···················· | ···················· | |
| D | 7449 hCoV-19/England/201140079/2020\|EPI_ISL_464714\|2020 | ···················· | ···················· | ···················· | ···················· | ···················· | ···················· | |
| D | 7452 hCoV-19/England/201140309/2020\|EPI_ISL_464721\|2020 | ···················· | ···················· | ···················· | ···················· | ···················· | ···················· | |
| D | 7453 hCoV-19/England/201140353/2020\|EPI_ISL_464726\|2020 | ···················· | ···················· | ···················· | ···················· | ··N·····NN·········· | ···················· | 1 |
| D | 7454 hCoV-19/England/201140359/2020\|EPI_ISL_464728\|2020 | ···················· | ···················· | ···················· | ···················· | ···················· | ···················· | |
| D | 7456 hCoV-19/England/201160010/2020\|EPI_ISL_464776\|2020 | ···················· | ···················· | ···················· | ···················· | ···················· | ···················· | |
| D | 7457 hCoV-19/England/201160011/2020\|EPI_ISL_464777\|2020 | ···················· | ···················· | ···················· | ···················· | ···················· | ···················· | |
| D | 7458 hCoV-19/England/201160012/2020\|EPI_ISL_464778\|2020 | ···················· | ···················· | ···················· | ···················· | ···················· | ···················· | |
| D | 7461 hCoV-19/England/201160049/2020\|EPI_ISL_464781\|2020 | ···················· | ···················· | ···················· | ···················· | ···················· | ···················· | |
| D | 7462 hCoV-19/England/201160064/2020\|EPI_ISL_464782\|2020 | ···················· | ···················· | ···················· | ···················· | ···················· | NN·················· | |
| D | 7463 hCoV-19/England/201160065/2020\|EPI_ISL_464783\|2020 | ···················· | ···················· | ···················· | ···················· | ···················· | ···················· | |
| D | 7464 hCoV-19/England/201160067/2020\|EPI_ISL_464784\|2020 | ···················· | ···················· | ···················· | ···················· | ···················· | ···················· | |
| D | 7465 hCoV-19/England/201160068/2020\|EPI_ISL_464785\|2020 | ···················· | ···················· | ···················· | ···················· | ···················· | ···················· | |
| D | 7467 hCoV-19/England/201160216/2020\|EPI_ISL_464788\|2020 | ···················· | ···················· | ···················· | ···················· | ···················· | ···················· | |
| D | 7469 hCoV-19/England/201160243/2020\|EPI_ISL_464790\|2020 | ···················· | ···················· | ···················· | ···················· | ···················· | ···················· | |
| D | 7470 hCoV-19/England/201160258/2020\|EPI_ISL_464791\|2020 | ···················· | ···················· | ···················· | ···················· | ···················· | N··N················ | |
| D | 7472 hCoV-19/England/201160271/2020\|EPI_ISL_464793\|2020 | ···················· | ···················· | ···················· | ···················· | ···················· | ···················· | |
| D | 7474 hCoV-19/England/201160276/2020\|EPI_ISL_464795\|2020 | ···················· | ···················· | ···················· | ···················· | ···················· | ···················· | |
| D | 7476 hCoV-19/England/201160628/2020\|EPI_ISL_464797\|2020 | ···················· | ···················· | ···················· | ···················· | ···················· | ···················· | |
| D | 7479 hCoV-19/Switzerland/AG-ETHZ-120002/2020\|EPI_ISL_46 | ···················· | ···················· | ···················· | ···················· | ···················· | ············R······· | |
| D | 7487 hCoV-19/Switzerland/ZH-ETHZ-120164/2020\|EPI_ISL_46 | ···················· | ···················· | ··········T········· | ···················· | ···················· | ···················· | |
| D | 7489 hCoV-19/Switzerland/BE-ETHZ-120169/2020\|EPI_ISL_46 | ···················· | ···················· | ···················· | ···················· | ···················· | ···················· | |
| D | 7492 hCoV-19/Switzerland/BS-ETHZ-120178/2020\|EPI_ISL_46 | ···················· | ···················· | ···················· | ···················· | ···················· | ···················· | |
| D | 7494 hCoV-19/Switzerland/BE-ETHZ-120182/2020\|EPI_ISL_46 | ···················· | ···················· | ···················· | ···················· | ···················· | ···················· | |
| D | 7498 hCoV-19/Switzerland/TI-ETHZ-120202/2020\|EPI_ISL_46 | ···················· | ···················· | ···················· | ···················· | ···················· | ···················· | |
| D | 7503 hCoV-19/Switzerland/BE-ETHZ-120221/2020\|EPI_ISL_46 | ···················· | ···················· | ···················· | ···················· | ···················· | ···················· | |
| D | 7505 hCoV-19/Switzerland/BE-ETHZ-120224/2020\|EPI_ISL_46 | ···················· | ···················· | ···················· | ···················· | ···················· | ···················· | |
| D | 7509 hCoV-19/England/BRIS-12CA0F/2020\|EPI_ISL_469504\|20 | ···················· | ···················· | ···················· | ···················· | ···················· | ···················· | |

| | | | | | | | | |
|---|---|---|---|---|---|---|---|---|
| D | 7510 | hCoV-19/England/BRIS-12C4E3/2020\|EPI_ISL_469505\|20 | ···················· | ···················· | ····················· | ···················· | ···················· | ···················· |
| D | 7511 | hCoV-19/England/BRIS-12C6FC/2020\|EPI_ISL_469506\|20 | ···················· | ···················· | ····················· | ···················· | ···················· | ···················· |
| D | 7512 | hCoV-19/England/BRIS-12C850/2020\|EPI_ISL_469507\|20 | ···················· | ···················· | ····················· | ···················· | ···················· | ···················· |
| D | 7513 | hCoV-19/England/BRIS-12C6DE/2020\|EPI_ISL_469508\|20 | ···················· | ···················· | ····················· | ···················· | ···················· | ···················· |
| D | 7514 | hCoV-19/England/BRIS-12C920/2020\|EPI_ISL_469509\|20 | ···················· | ···················· | ····················· | ···················· | ···················· | ···················· |
| D | 7515 | hCoV-19/England/BRIS-12C7DB/2020\|EPI_ISL_469510\|20 | ···················· | ···················· | ····················· | ···················· | ···················· | ···················· |
| D | 7516 | hCoV-19/England/BRIS-12C5B3/2020\|EPI_ISL_469513\|20 | ···················· | ···················· | ····················· | ···················· | ···················· | ···················· |
| D | 7517 | hCoV-19/England/BRIS-12C95D/2020\|EPI_ISL_469515\|20 | ···················· | ···················· | ····················· | ···················· | ···················· | ···················· |
| D | 7518 | hCoV-19/England/BRIS-12C911/2020\|EPI_ISL_469516\|20 | ···················· | ···················· | ····················· | ···················· | ···················· | ···················· |
| D | 7519 | hCoV-19/England/BRIS-12C5A4/2020\|EPI_ISL_469517\|20 | ···················· | ···················· | ····················· | ···················· | ···················· | ···················· |
| D | 7520 | hCoV-19/England/BRIS-12C7CC/2020\|EPI_ISL_469518\|20 | ···················· | ···················· | ····················· | ···················· | ···················· | ···················· |
| D | 7521 | hCoV-19/England/BRIS-12C53B/2020\|EPI_ISL_469519\|20 | ···················· | ···················· | ····················· | ···················· | ···················· | ···················· |
| D | 7522 | hCoV-19/England/BRIS-12C559/2020\|EPI_ISL_469520\|20 | ···················· | ···················· | ····················· | ···················· | ···················· | ···················· |
| D | 7523 | hCoV-19/England/BRIS-12C9B7/2020\|EPI_ISL_469521\|20 | ···················· | ···················· | ····················· | ···················· | ···················· | ···················· |
| D | 7524 | hCoV-19/England/BRIS-12C5D1/2020\|EPI_ISL_469523\|20 | ···················· | ···················· | ····················· | ···················· | ···················· | ···················· |
| D | 7525 | hCoV-19/England/BRIS-12C665/2020\|EPI_ISL_469524\|20 | ···················· | ···················· | ····················· | ···················· | ···················· | ···················· |
| D | 7526 | hCoV-19/England/BRIS-12C735/2020\|EPI_ISL_469525\|20 | ···················· | ···················· | ····················· | ···················· | ···················· | ···················· |
| D | 7527 | hCoV-19/England/BRIS-131885/2020\|EPI_ISL_469526\|20 | ···················· | ···················· | ····················· | ···················· | ···················· | ···················· |
| D | 7528 | hCoV-19/England/BRIS-127521/2020\|EPI_ISL_469529\|20 | ···················· | ···················· | ····················· | ···················· | ···················· | ···················· |
| D | 7529 | hCoV-19/England/BRIS-1282A1/2020\|EPI_ISL_469531\|20 | ···················· | ···················· | ····················· | ···················· | ···················· | ···················· |
| D | 7530 | hCoV-19/England/BRIS-1271F0/2020\|EPI_ISL_469532\|20 | ···················· | ···················· | ····················· | ···················· | ···················· | ···················· |
| D | 7531 | hCoV-19/England/BRIS-126EBE/2020\|EPI_ISL_469535\|20 | ···················· | ···················· | ····················· | ···················· | ···················· | ···················· |
| D | 7532 | hCoV-19/England/BRIS-127020/2020\|EPI_ISL_469536\|20 | ···················· | ···A················ | ····················· | ···················· | ···················· | ···················· |
| D | 7533 | hCoV-19/England/BRIS-128441/2020\|EPI_ISL_469537\|20 | ···················· | ···················· | ····················· | ···················· | ···················· | ···················· |
| D | 7534 | hCoV-19/England/BRIS-127FD8/2020\|EPI_ISL_469538\|20 | ···················· | ···················· | ····················· | ···················· | ···················· | ···················· |
| D | 7535 | hCoV-19/England/BRIS-128010/2020\|EPI_ISL_469539\|20 | ···················· | ···················· | ····················· | ···················· | ···················· | ···················· |
| D | 7536 | hCoV-19/England/BRIS-126EDC/2020\|EPI_ISL_469543\|20 | ···················· | ···················· | ····················· | ···················· | ···················· | ···················· |
| D | 7537 | hCoV-19/England/BRIS-127C4A/2020\|EPI_ISL_469544\|20 | ···················· | ···················· | ····················· | ···················· | ···················· | ···················· |
| D | 7538 | hCoV-19/England/BRIS-1284C9/2020\|EPI_ISL_469546\|20 | ···················· | ···················· | ····················· | ···················· | ···················· | ···················· |
| D | 7539 | hCoV-19/England/BRIS-12708A/2020\|EPI_ISL_469547\|20 | ···················· | ···················· | ····················· | ···················· | ···················· | ···················· |
| D | 7540 | hCoV-19/England/BRIS-127C3B/2020\|EPI_ISL_469562\|20 | ···················· | ···················· | ····················· | ···················· | ···················· | ···················· |
| D | 7541 | hCoV-19/England/BRIS-1283EA/2020\|EPI_ISL_469564\|20 | ···················· | ···················· | ····················· | ···················· | ···················· | ···················· |
| D | 7542 | hCoV-19/England/BRIS-12814A/2020\|EPI_ISL_469570\|20 | ···················· | ···················· | ····················· | ···················· | ···················· | ···················· |
| D | 7543 | hCoV-19/England/BRIS-128423/2020\|EPI_ISL_469571\|20 | ···················· | ···················· | ····················· | ···················· | ···················· | ···················· |
| D | 7544 | hCoV-19/England/BRIS-1278DD/2020\|EPI_ISL_469573\|20 | ···················· | ···················· | ····················· | ···················· | ···················· | ···················· |
| D | 7545 | hCoV-19/England/BRIS-127363/2020\|EPI_ISL_469574\|20 | ···················· | ···················· | ····················· | ···················· | ···················· | ···················· |
| D | 7546 | hCoV-19/England/BRIS-12758B/2020\|EPI_ISL_469577\|20 | ···················· | ···················· | ····················· | ···················· | ···················· | ···················· |
| D | 7547 | hCoV-19/England/BRIS-1273AF/2020\|EPI_ISL_469579\|20 | ···················· | ···················· | ····················· | ···················· | ···················· | ···················· |
| D | 7548 | hCoV-19/England/BRIS-127187/2020\|EPI_ISL_469580\|20 | ···················· | ···················· | ····················· | ···················· | ···················· | ···················· |
| D | 7549 | hCoV-19/England/BRIS-128177/2020\|EPI_ISL_469581\|20 | ···················· | ···················· | ····················· | ···················· | ···················· | ···················· |
| D | 7550 | hCoV-19/England/BRIS-127C95/2020\|EPI_ISL_469582\|20 | ···················· | ···················· | ····················· | ···················· | ···················· | ···················· |
| D | 7551 | hCoV-19/England/BRIS-1270E4/2020\|EPI_ISL_469583\|20 | ···················· | ···················· | ····················· | ···················· | ···················· | ···················· |
| D | 7552 | hCoV-19/England/BRIS-126FE8/2020\|EPI_ISL_469584\|20 | ···················· | ···················· | ····················· | ···················· | ···················· | ···················· |
| D | 7553 | hCoV-19/England/BRIS-127390/2020\|EPI_ISL_469586\|20 | ···················· | ···················· | ····················· | ···················· | ···················· | ···················· |
| D | 7554 | hCoV-19/England/BRIS-12710F/2020\|EPI_ISL_469588\|20 | ···················· | ···················· | ····················· | ···················· | ···················· | ···················· |
| D | 7555 | hCoV-19/England/BRIS-127F7E/2020\|EPI_ISL_469591\|20 | ···················· | ···················· | ····················· | ···················· | ···················· | ···················· |
| D | 7556 | hCoV-19/England/CAMB-800CC/2020\|EPI_ISL_470500\|202 | ···················· | ···················· | ····················· | ···················· | ···················· | ···················· |
| D | 7557 | hCoV-19/England/CAMB-801BA/2020\|EPI_ISL_470501\|202 | ···················· | ···················· | ····················· | ···················· | ···················· | ···················· |
| D | 7558 | hCoV-19/England/CAMB-80572/2020\|EPI_ISL_470502\|202 | ···················· | ···················· | ····················· | ···················· | ···················· | ···················· |
| D | 7559 | hCoV-19/England/CAMB-809A3/2020\|EPI_ISL_470503\|202 | ···················· | ···················· | ····················· | ···················· | ···················· | ···················· |
| D | 7560 | hCoV-19/England/CAMB-80633/2020\|EPI_ISL_470504\|202 | ···················· | ···················· | ····················· | ···················· | ···················· | ···················· |
| D | 7561 | hCoV-19/England/CAMB-7FA4A/2020\|EPI_ISL_470505\|202 | ···················· | ···················· | ····················· | ···················· | ···················· | ···················· |
| D | 7562 | hCoV-19/England/CAMB-809C1/2020\|EPI_ISL_470506\|202 | ···················· | ···················· | ····················· | ···················· | ···················· | ···················· |
| D | 7563 | hCoV-19/England/CAMB-8016F/2020\|EPI_ISL_470507\|202 | ···················· | ···················· | ····················· | ···················· | ···················· | ···················· |
| D | 7564 | hCoV-19/England/CAMB-80ABF/2020\|EPI_ISL_470508\|202 | ···················· | ···················· | ····················· | ···················· | ···················· | ···················· |
| D | 7565 | hCoV-19/England/CAMB-7F813/2020\|EPI_ISL_470510\|202 | ···················· | ···················· | ····················· | ···················· | ···················· | ···················· |
| D | 7566 | hCoV-19/England/CAMB-7F910/2020\|EPI_ISL_470511\|202 | ···················· | ·A·················· | ····················· | ···················· | ···················· | ···················· |
| D | 7567 | hCoV-19/England/CAMB-80123/2020\|EPI_ISL_470512\|202 | ···················· | ···················· | ····················· | ···················· | ···················· | ···················· |
| D | 7568 | hCoV-19/England/BRIS-1237E3/2020\|EPI_ISL_470530\|20 | ···················· | ···················· | ····················· | ···················· | ···················· | ···················· |
| D | 7569 | hCoV-19/England/CAMB-7C88F/2020\|EPI_ISL_470531\|202 | ···················· | ···················· | ····················· | ···················· | ···················· | ···················· |
| D | 7570 | hCoV-19/England/CAMB-7CBB3/2020\|EPI_ISL_470532\|202 | ···················· | ···················· | ····················· | ···················· | ···················· | ···················· |
| D | 7571 | hCoV-19/England/CAMB-7CE32/2020\|EPI_ISL_470533\|202 | ···················· | ···················· | ····················· | ···················· | ···················· | ···················· |
| D | 7572 | hCoV-19/England/CAMB-78FCA/2020\|EPI_ISL_470535\|202 | ···················· | ···················· | ····················· | ···················· | ···················· | ···················· |
| D | 7573 | hCoV-19/England/CAMB-79318/2020\|EPI_ISL_470536\|202 | ···················· | ···················· | ····················· | ···················· | ···················· | ···················· |
| D | 7574 | hCoV-19/England/CAMB-78E63/2020\|EPI_ISL_470537\|202 | ···················· | ···················· | ····················· | ···················· | ···················· | ···················· |
| D | 7575 | hCoV-19/Russia/CRIE130014/2020\|EPI_ISL_470539\|2020 | ···················· | ···················· | ····················· | ···················· | ···················· | ···················· |
| D | 7576 | hCoV-19/USA/WI-WSLH-200011/2020\|EPI_ISL_470540\|202 | ···················· | ···················· | ····················· | ···················· | ···················· | ···················· |
| D | 7577 | hCoV-19/USA/UT-01923/2020\|EPI_ISL_470544\|2020-04-1 | ···················· | ···················· | ····················· | ···················· | ···················· | ···················· |
| D | 7578 | hCoV-19/USA/UT-02432/2020\|EPI_ISL_470549\|2020-04-2 | ···················· | ···················· | ····················· | ···················· | ···················· | ···················· |
| D | 7579 | hCoV-19/USA/UT-02434/2020\|EPI_ISL_470550\|2020-04-2 | ···················· | ···················· | ····················· | ···················· | ···················· | ···················· |
| D | 7580 | hCoV-19/USA/UT-02435/2020\|EPI_ISL_470551\|2020-04-2 | ···················· | ···················· | ····················· | ···················· | ···················· | ···················· |
| D | 7581 | hCoV-19/USA/UT-02437/2020\|EPI_ISL_470552\|2020-04-2 | ···················· | ···················· | ····················· | ···················· | ···················· | ···················· |
| D | 7582 | hCoV-19/USA/UT-02438/2020\|EPI_ISL_470553\|2020-04-2 | ···················· | ···················· | ····················· | ···················· | ···················· | ···················· |
| D | 7583 | hCoV-19/USA/UT-03892/2020\|EPI_ISL_470554\|2020-04-1 | ···················· | ···················· | ····················· | ···················· | ···················· | ···················· |
| D | 7584 | hCoV-19/USA/UT-03901/2020\|EPI_ISL_470559\|2020-04-1 | ···················· | ···················· | ····················· | ···················· | ···················· | ···················· |
| D | 7585 | hCoV-19/USA/UT-03966/2020\|EPI_ISL_470565\|2020-04-2 | ···················· | ···················· | ····················· | ···················· | ···················· | ···················· |
| D | 7586 | hCoV-19/USA/UT-04836/2020\|EPI_ISL_470566\|2020-04-1 | ···················· | ···················· | ····················· | ···················· | ···················· | ···················· |
| D | 7587 | hCoV-19/USA/MI-MDHHS-SC20799/2020\|EPI_ISL_471820\|2 | ···················· | ···················· | ····················· | ···················· | ···················· | ···················· |

| | | | | | | | | |
|---|---|---|---|---|---|---|---|---|
| D | 7588 | hCoV-19/USA/MI-MDHHS-SC20800/2020\|EPI_ISL_471821\|2 | ···················· | ···················· | ····················· | ···················· | ···················· | ···················· |
| D | 7589 | hCoV-19/USA/MI-MDHHS-SC20801/2020\|EPI_ISL_471822\|2 | ···················· | ···················· | ····················· | ···················· | ···················· | ···················· |
| D | 7590 | hCoV-19/USA/MI-MDHHS-SC20802/2020\|EPI_ISL_471823\|2 | ···················· | ···················· | ····················· | ···················· | ···················· | ···················· |
| D | 7591 | hCoV-19/USA/MI-MDHHS-SC20804/2020\|EPI_ISL_471825\|2 | ···················· | ···················· | ····················· | ···················· | ···················· | ···················· |
| D | 7592 | hCoV-19/USA/MI-MDHHS-SC20805/2020\|EPI_ISL_471826\|2 | ···················· | ···················· | ····················· | ···················· | ···················· | ···················· |
| D | 7593 | hCoV-19/USA/MI-MDHHS-SC20809/2020\|EPI_ISL_471830\|2 | ···················· | ···················· | ····················· | ···················· | ···················· | ···················· |
| D | 7594 | hCoV-19/USA/MI-MDHHS-SC20810/2020\|EPI_ISL_471831\|2 | ···················· | ···················· | ····················· | ···················· | ···················· | ···················· |
| D | 7595 | hCoV-19/USA/MI-MDHHS-SC20811/2020\|EPI_ISL_471832\|2 | ···················· | ···················· | ····················· | ···················· | ···················· | ···················· |
| D | 7596 | hCoV-19/USA/MI-MDHHS-SC20812/2020\|EPI_ISL_471833\|2 | ···················· | ···················· | ····················· | ···················· | ···················· | ···················· |
| D | 7597 | hCoV-19/USA/MI-MDHHS-SC20813/2020\|EPI_ISL_471834\|2 | ···················· | ···················· | ····················· | ···················· | ···················· | ···················· |
| D | 7598 | hCoV-19/USA/MI-MDHHS-SC20814/2020\|EPI_ISL_471835\|2 | ···················· | ···················· | ····················· | ···················· | ···················· | ···················· |
| D | 7599 | hCoV-19/USA/MI-MDHHS-SC20816/2020\|EPI_ISL_471837\|2 | ···················· | ···················· | ····················· | ···················· | ···················· | ···················· |
| D | 7600 | hCoV-19/USA/MI-MDHHS-SC20817/2020\|EPI_ISL_471838\|2 | ···················· | ···················· | ····················· | ···················· | ···················· | ···················· |
| D | 7601 | hCoV-19/USA/MI-MDHHS-SC20818/2020\|EPI_ISL_471839\|2 | ···················· | ···················· | ····················· | ···················· | ···················· | ···················· |
| D | 7602 | hCoV-19/USA/MI-MDHHS-SC20819/2020\|EPI_ISL_471840\|2 | ···················· | ···················· | ····················· | ···················· | ···················· | ···················· |
| D | 7603 | hCoV-19/USA/MI-MDHHS-SC20821/2020\|EPI_ISL_471842\|2 | ···················· | ···················· | ····················· | ···················· | ···················· | ···················· |
| D | 7604 | hCoV-19/USA/MI-MDHHS-SC20822/2020\|EPI_ISL_471843\|2 | ···················· | ··C················· | ····················· | ···················· | ···················· | ···················· |
| D | 7605 | hCoV-19/USA/MI-MDHHS-SC20850/2020\|EPI_ISL_471871\|2 | ···················· | ···················· | ····················· | ···················· | ···················· | ···················· |
| D | 7606 | hCoV-19/USA/MI-MDHHS-SC20851/2020\|EPI_ISL_471872\|2 | ···················· | ···················· | ····················· | ···················· | ···················· | ···················· |
| D | 7607 | hCoV-19/USA/MI-MDHHS-SC20852/2020\|EPI_ISL_471873\|2 | ···················· | ···················· | ····················· | ···················· | ···················· | ···················· |
| D | 7608 | hCoV-19/USA/MI-MDHHS-SC20853/2020\|EPI_ISL_471874\|2 | ···················· | ···················· | ····················· | ···················· | ···················· | ···················· |
| D | 7609 | hCoV-19/USA/MI-MDHHS-SC20854/2020\|EPI_ISL_471875\|2 | ···················· | ···················· | ····················· | ···················· | ···················· | ···················· |
| D | 7610 | hCoV-19/USA/MI-MDHHS-SC20855/2020\|EPI_ISL_471876\|2 | ···················· | ···················· | ····················· | ···················· | ···················· | ···················· |
| D | 7611 | hCoV-19/USA/MI-MDHHS-SC20856/2020\|EPI_ISL_471877\|2 | ···················· | ···················· | ····················· | ···················· | ···················· | ···················· |
| D | 7612 | hCoV-19/USA/MI-MDHHS-SC20857/2020\|EPI_ISL_471878\|2 | ···················· | ···················· | ····················· | ···················· | ···················· | ···················· |
| D | 7613 | hCoV-19/USA/MI-MDHHS-SC20858/2020\|EPI_ISL_471879\|2 | ···················· | ···················· | ····················· | ···················· | ···················· | ···················· |
| D | 7614 | hCoV-19/USA/MI-MDHHS-SC20859/2020\|EPI_ISL_471880\|2 | ···················· | ···················· | ····················· | ···················· | ···················· | ···················· |
| D | 7615 | hCoV-19/USA/MI-MDHHS-SC20860/2020\|EPI_ISL_471881\|2 | ···················· | ···················· | ····················· | ···················· | ···················· | ···················· |
| D | 7616 | hCoV-19/USA/MI-MDHHS-SC20861/2020\|EPI_ISL_471882\|2 | ···················· | ···················· | ····················· | ···················· | ···················· | ···················· |
| D | 7617 | hCoV-19/USA/MI-MDHHS-SC20862/2020\|EPI_ISL_471883\|2 | ···················· | ···················· | ····················· | ···················· | ···················· | ···················· |
| D | 7618 | hCoV-19/USA/MI-MDHHS-SC20863/2020\|EPI_ISL_471884\|2 | ···················· | ···················· | ····················· | ···················· | ···················· | ···················· |
| D | 7619 | hCoV-19/USA/MI-MDHHS-SC20864/2020\|EPI_ISL_471885\|2 | ···················· | ···················· | ····················· | ···················· | ···················· | ···················· |
| D | 7620 | hCoV-19/USA/MI-MDHHS-SC20865/2020\|EPI_ISL_471886\|2 | ···················· | ···················· | ····················· | ···················· | ···················· | ···················· |
| D | 7621 | hCoV-19/USA/MI-MDHHS-SC20866/2020\|EPI_ISL_471887\|2 | ···················· | ···················· | ····················· | ···················· | ···················· | ···················· |
| D | 7622 | hCoV-19/USA/MI-MDHHS-SC20867/2020\|EPI_ISL_471888\|2 | ···················· | ···················· | ····················· | ···················· | ···················· | ···················· |
| D | 7623 | hCoV-19/USA/MI-MDHHS-SC20868/2020\|EPI_ISL_471889\|2 | ···················· | ···················· | ····················· | ···················· | ···················· | ···················· |
| D | 7624 | hCoV-19/USA/MI-MDHHS-SC20869/2020\|EPI_ISL_471890\|2 | ···················· | ···················· | ····················· | ···················· | ···················· | ···················· |
| D | 7625 | hCoV-19/USA/MI-MDHHS-SC20870/2020\|EPI_ISL_471891\|2 | ···················· | ···················· | ····················· | ···················· | ···················· | ···················· |
| D | 7626 | hCoV-19/USA/MI-MDHHS-SC20875/2020\|EPI_ISL_471896\|2 | ···················· | ···················· | ····················· | ···················· | ···················· | ···················· |
| D | 7627 | hCoV-19/USA/MI-MDHHS-SC20876/2020\|EPI_ISL_471897\|2 | ···················· | ···················· | ····················· | ···················· | ···················· | ···················· |
| D | 7628 | hCoV-19/USA/MI-MDHHS-SC20877/2020\|EPI_ISL_471898\|2 | ···················· | ···················· | ····················· | ···················· | ···················· | ···················· |
| D | 7629 | hCoV-19/USA/MI-MDHHS-SC20878/2020\|EPI_ISL_471899\|2 | ···················· | ···················· | ····················· | ···················· | ···················· | ···················· |
| D | 7630 | hCoV-19/England/SHEF-C7ADA/2020\|EPI_ISL_475401\|202 | ···················· | ···················· | ····················· | ···················· | ···················· | ···················· |
| D | 7631 | hCoV-19/England/SHEF-C7E1A/2020\|EPI_ISL_475402\|202 | ···················· | ···················· | ····················· | ···················· | ···················· | ···················· |
| D | 7632 | hCoV-19/England/SHEF-C7E29/2020\|EPI_ISL_475403\|202 | ···················· | ···················· | ····················· | ···················· | ···················· | ···················· |
| D | 7633 | hCoV-19/England/SHEF-C86A8/2020\|EPI_ISL_475407\|202 | ···················· | ···················· | ····················· | ···················· | ···················· | ···················· |
| D | 7634 | hCoV-19/England/SHEF-C870F/2020\|EPI_ISL_475409\|202 | ···················· | ···················· | ····················· | ···················· | ···················· | ···················· |
| D | 7635 | hCoV-19/England/SHEF-C8EEC/2020\|EPI_ISL_475413\|202 | ···················· | ···················· | ····················· | ···················· | ···················· | ···················· |
| D | 7636 | hCoV-19/England/SHEF-C8FBC/2020\|EPI_ISL_475415\|202 | ···················· | ···················· | ····················· | ···················· | ···················· | ···················· |
| D | 7637 | hCoV-19/England/SHEF-C91E2/2020\|EPI_ISL_475417\|202 | ···················· | ···················· | ····················· | ···················· | ···················· | ···················· |
| D | 7638 | hCoV-19/England/SHEF-C9267/2020\|EPI_ISL_475418\|202 | ···················· | ···················· | ····················· | ···················· | ···················· | ···················· |
| D | 7639 | hCoV-19/England/SHEF-C9319/2020\|EPI_ISL_475419\|202 | ···················· | ···················· | ····················· | ···················· | ···················· | ···················· |
| D | 7640 | hCoV-19/England/SHEF-C9470/2020\|EPI_ISL_475420\|202 | ···················· | ···················· | ····················· | ···················· | ···················· | ···················· |
| D | 7641 | hCoV-19/England/SHEF-C948F/2020\|EPI_ISL_475421\|202 | ···················· | ···················· | ····················· | ···················· | ···················· | ···················· |
| D | 7642 | hCoV-19/Venezuela/VEN-89312/2020\|EPI_ISL_476702\|20 | ···················· | ···················· | ····················· | ···················· | ···················· | ···················· |
| D | 7643 | hCoV-19/Venezuela/VEN-95072/2020\|EPI_ISL_476704\|20 | ···················· | ···················· | ····················· | ···················· | ···················· | ···················· |
| D | 7644 | hCoV-19/Germany/NW-MPP-1/2020\|EPI_ISL_476705\|2020- | ···················· | ···················· | ····················· | ···················· | ···················· | ···················· |
| D | 7645 | hCoV-19/USA/MN-MDH-1121/2020\|EPI_ISL_476706\|2020-0 | ···················· | ···················· | ····················· | ···················· | ···················· | ···················· |
| D | 7646 | hCoV-19/USA/MN-MDH-1122/2020\|EPI_ISL_476707\|2020-0 | ···················· | ···················· | ····················· | ···················· | ···················· | ···················· |
| D | 7647 | hCoV-19/USA/MN-MDH-1123/2020\|EPI_ISL_476708\|2020-0 | ···················· | ···················· | ····················· | ···················· | ···················· | ···················· |
| D | 7648 | hCoV-19/USA/MN-MDH-1124/2020\|EPI_ISL_476709\|2020-0 | ···················· | ···················· | ····················· | ···················· | ···················· | ···················· |
| D | 7649 | hCoV-19/USA/MN-MDH-1135/2020\|EPI_ISL_476720\|2020-0 | ···················· | ···················· | ····················· | ···················· | ···················· | ···················· |
| D | 7650 | hCoV-19/USA/MN-MDH-1136/2020\|EPI_ISL_476721\|2020-0 | ···················· | ···················· | ····················· | ···················· | ···················· | ···················· |
| D | 7651 | hCoV-19/USA/MN-MDH-1137/2020\|EPI_ISL_476722\|2020-0 | ···················· | ···················· | ····················· | ···················· | ···················· | ···················· |
| D | 7652 | hCoV-19/USA/MN-MDH-1138/2020\|EPI_ISL_476723\|2020-0 | ···················· | ···················· | ····················· | ···················· | ···················· | ···················· |
| D | 7653 | hCoV-19/USA/MN-MDH-1139/2020\|EPI_ISL_476724\|2020-0 | ···················· | ···················· | ····················· | ···················· | ···················· | ···················· |
| D | 7654 | hCoV-19/USA/MN-MDH-1140/2020\|EPI_ISL_476725\|2020-0 | ···················· | ···················· | ····················· | ···················· | ···················· | ···················· |
| D | 7655 | hCoV-19/USA/MN-MDH-1142/2020\|EPI_ISL_476727\|2020-0 | ···················· | ···················· | ····················· | ···················· | ···················· | ···················· |
| D | 7656 | hCoV-19/USA/MN-MDH-1143/2020\|EPI_ISL_476728\|2020-0 | ···················· | ···················· | ····················· | ···················· | ···················· | ···················· |
| D | 7657 | hCoV-19/USA/MN-MDH-1144/2020\|EPI_ISL_476729\|2020-0 | ···················· | ···················· | ····················· | ···················· | ···················· | ···················· |
| D | 7658 | hCoV-19/USA/MN-MDH-1145/2020\|EPI_ISL_476730\|2020-0 | ···················· | ···················· | ····················· | ···················· | ···················· | ···················· |
| D | 7659 | hCoV-19/USA/MN-MDH-1146/2020\|EPI_ISL_476731\|2020-0 | ···················· | ···················· | ····················· | ···················· | ···················· | ···················· |
| D | 7660 | hCoV-19/USA/MN-MDH-1147/2020\|EPI_ISL_476732\|2020-0 | ···················· | ···················· | ····················· | ···················· | ···················· | ···················· |
| D | 7661 | hCoV-19/USA/MN-MDH-1148/2020\|EPI_ISL_476733\|2020-0 | ···················· | ···················· | ····················· | ···················· | ···················· | ···················· |
| D | 7662 | hCoV-19/USA/MN-MDH-1149/2020\|EPI_ISL_476734\|2020-0 | ···················· | ···················· | ····················· | ···················· | ···················· | ···················· |
| D | 7663 | hCoV-19/USA/MN-MDH-1150/2020\|EPI_ISL_476735\|2020-0 | ···················· | ···················· | ····················· | ···················· | ···················· | ···················· |
| D | 7664 | hCoV-19/USA/MN-MDH-1151/2020\|EPI_ISL_476736\|2020-0 | ···················· | ···················· | ····················· | ···················· | ···················· | ···················· |
| D | 7665 | hCoV-19/USA/MN-MDH-1152/2020\|EPI_ISL_476737\|2020-0 | ···················· | ···················· | ····················· | ···················· | ···················· | ···················· |

D 7666 hCoV-19/USA/MN-MDH-1153/2020|EPI_ISL_476738|2020-0 .................... .................... .................... .................... .................... ....................
D 7667 hCoV-19/USA/MN-MDH-1154/2020|EPI_ISL_476739|2020-0 .................... .................... .................... .................... .................... ....................
D 7668 hCoV-19/USA/MN-MDH-1155/2020|EPI_ISL_476740|2020-0 .................... .................... .................... .................... .................... ....................
D 7669 hCoV-19/USA/MN-MDH-1156/2020|EPI_ISL_476741|2020-0 .................... .................... .................... .................... .................... ....................
D 7670 hCoV-19/USA/MN-MDH-1157/2020|EPI_ISL_476742|2020-0 .................... .................... .................... .................... .................... ....................
D 7671 hCoV-19/USA/MN-MDH-1158/2020|EPI_ISL_476743|2020-0 .................... .................... .................... .................... .................... ....................
D 7672 hCoV-19/USA/MN-MDH-1159/2020|EPI_ISL_476744|2020-0 .................... .................... .................... .................... .................... ....................
D 7673 hCoV-19/USA/MN-MDH-1160/2020|EPI_ISL_476745|2020-0 .................... .................... .................... .................... .................... ....................
D 7674 hCoV-19/USA/MN-MDH-1161/2020|EPI_ISL_476746|2020-0 .................... .................... .................... .................... .................... ....................
D 7675 hCoV-19/USA/MN-MDH-1162/2020|EPI_ISL_476747|2020-0 .................... .................... .................... .................... .................... ....................
D 7676 hCoV-19/USA/MN-MDH-1163/2020|EPI_ISL_476748|2020-0 .................... .................... .................... .................... .................... ....................
D 7677 hCoV-19/USA/MN-MDH-1164/2020|EPI_ISL_476749|2020-0 .................... .................... .................... .................... .................... ....................
D 7678 hCoV-19/USA/MN-MDH-1165/2020|EPI_ISL_476750|2020-0 .................... .................... .................... .................... .................... ....................
D 7679 hCoV-19/USA/MN-MDH-1166/2020|EPI_ISL_476751|2020-0 .................... .................... .................... .................... .................... ....................
D 7680 hCoV-19/USA/MN-MDH-1167/2020|EPI_ISL_476752|2020-0 .................... .................... .................... .................... .................... ....................
D 7681 hCoV-19/Belgium/VHEJ-0329357/2020|EPI_ISL_462211|2 .................... .................... .................... .................... .................... ....................
D 7682 hCoV-19/Belgium/TA-0330358/2020|EPI_ISL_462212|202 .................... .................... .................... .................... .................... ....................
D 7683 hCoV-19/Belgium/BS-0331359/2020|EPI_ISL_462213|202 .................... .................... .................... .................... .................... ....................
D 7684 hCoV-19/Belgium/SN-0331360/2020|EPI_ISL_462214|202 .................... .................... .................... .................... .................... ....................
D 7685 hCoV-19/Belgium/BS-0331361/2020|EPI_ISL_462215|202 .................... .................... .................... .................... .................... ....................
D 7686 hCoV-19/Belgium/BD0331362/2020|EPI_ISL_462216|2020 .................... .................... .................... .................... .................... ....................
D 7687 hCoV-19/Belgium/MFM-0330363/2020|EPI_ISL_462217|20 .................... .................... .................... .................... .................... ....................
D 7688 hCoV-19/Belgium/LPJ-0401364/2020|EPI_ISL_462218|20 .................... .................... .................... .................... .................... ....................
D 7689 hCoV-19/Belgium/PBT-0330365/2020|EPI_ISL_462219|20 .................... .................... .................... .................... .................... ....................
D 7690 hCoV-19/Belgium/DRR-0401366/2020|EPI_ISL_462220|20 .................... .................... .................... .................... .................... ....................
D 7691 hCoV-19/Belgium/SAA-0330367/2020|EPI_ISL_462221|20 .................... .................... .................... .................... .................... ....................
D 7692 hCoV-19/Belgium/LJA-0401368/2020|EPI_ISL_462222|20 .................... .................... .................... .................... .................... ....................
D 7693 hCoV-19/Belgium/DAA-0327369/2020|EPI_ISL_462223|20 .................... .................... .................... .................... .................... ....................
D 7694 hCoV-19/Belgium/VAJ-0328370/2020|EPI_ISL_462224|20 .................... .................... .................... .................... .................... ....................
D 7695 hCoV-19/Belgium/VBMJ-0401430/2020|EPI_ISL_462225|2 .................... .................... .................... .................... .................... ....................
D 7696 hCoV-19/Belgium/VRMJ-0401372/2020|EPI_ISL_462226|2 .................... .................... .................... .................... .................... ....................
D 7697 hCoV-19/Belgium/IMJ-0401373/2020|EPI_ISL_462227|20 .................... .................... .................... .................... .................... ....................
D 7698 hCoV-19/Belgium/VGM-0402374/2020|EPI_ISL_462228|20 .................... .................... .................... .................... .................... ....................
D 7699 hCoV-19/Belgium/OTG-0401375/2020|EPI_ISL_462229|20 .................... .................... .................... .................... .................... ....................
D 7700 hCoV-19/Belgium/HHC-0401376/2020|EPI_ISL_462230|20 .................... .................... .................... .................... .................... ....................
D 7701 hCoV-19/Belgium/LL-0331377/2020|EPI_ISL_462231|202 .................... .................... .................... .................... .................... ....................
D 7702 hCoV-19/Belgium/SC-0331378/2020|EPI_ISL_462232|202 .................... .................... .................... .................... .................... ....................
D 7703 hCoV-19/Singapore/159/2020|EPI_ISL_462277|2020-03- .................... .................... .................... .................... .................... ....................
D 7704 hCoV-19/Singapore/160/2020|EPI_ISL_462278|2020-03- .................... .................... .................... .................... .................... ....................
D 7705 hCoV-19/Singapore/161/2020|EPI_ISL_462279|2020-03- .................... .................... .................... .................... .................... ....................
D 7706 hCoV-19/Singapore/162/2020|EPI_ISL_462280|2020-03- .................... .................... .................... .................... .................... ....................
D 7707 hCoV-19/Singapore/163/2020|EPI_ISL_462281|2020-03- .................... .................... .................... .................... .................... ....................
D 7708 hCoV-19/Singapore/164/2020|EPI_ISL_462282|2020-03- .................... .................... .................... .................... .................... ....................
D 7709 hCoV-19/Singapore/165/2020|EPI_ISL_462283|2020-01- .................... .................... .................... .................... .................... ....................
D 7710 hCoV-19/Singapore/166/2020|EPI_ISL_462284|2020-03- .................... .................... .................... .................... .................... ....................
D 7711 hCoV-19/Singapore/167/2020|EPI_ISL_462285|2020-03- .................... .................... .................... .................... .................... ....................
D 7712 hCoV-19/Singapore/168/2020|EPI_ISL_462286|2020-03- .................... .................... .................... .................... .................... ....................
D 7713 hCoV-19/Singapore/169/2020|EPI_ISL_462287|2020-03- .................... .................... .................... .................... .................... ...................R 1
D 7714 hCoV-19/Singapore/170/2020|EPI_ISL_462288|2020-04- .................... .................... .................... .................... .................... ....................
D 7715 hCoV-19/Singapore/172/2020|EPI_ISL_462290|2020-03- .................... .................... .................... .................... .................... ....................
D 7716 hCoV-19/Singapore/173/2020|EPI_ISL_462291|2020-03- .................... .................... .................... .................... .................... ....................
D 7717 hCoV-19/Singapore/174/2020|EPI_ISL_462292|2020-03- .................... .................... .................... .................... .................... ....................
D 7718 hCoV-19/Singapore/175/2020|EPI_ISL_462293|2020-02- .................... .................... .................... .................... .................... ....................
D 7719 hCoV-19/Singapore/177/2020|EPI_ISL_462295|2020-03- .................... .................... .................... .................... .................... ....................
D 7720 hCoV-19/Singapore/178/2020|EPI_ISL_462296|2020-03- .................... .................... .................... .................... .................... ....................
D 7721 hCoV-19/Singapore/179/2020|EPI_ISL_462297|2020-03- .................... .................... .................... .................... .................... ....................
D 7722 hCoV-19/Singapore/180/2020|EPI_ISL_462298|2020-03- .................... .................... .................... .................... .................... ....................
D 7723 hCoV-19/Singapore/181/2020|EPI_ISL_462299|2020-03- .................... .................... .................... .................... .................... ....................
D 7724 hCoV-19/USA/WA-S1052/2020|EPI_ISL_463542|2020-05-0 .................... .................... .................... .................... .................... ....................
D 7725 hCoV-19/USA/WA-S1053/2020|EPI_ISL_463543|2020-05-0 .................... .................... .................... .................... .................... ....................
D 7726 hCoV-19/USA/WA-S1054/2020|EPI_ISL_463544|2020-04-3 .................... .................... .................... .................... .................... ....................
D 7727 hCoV-19/USA/WA-S1055/2020|EPI_ISL_463545|2020-04-3 .................... .................... .................... .................... .................... ....................
D 7728 hCoV-19/USA/WA-S1056/2020|EPI_ISL_463546|2020-04-3 .................... .................... .................... .................... .................... ....................
D 7729 hCoV-19/USA/WA-S1057/2020|EPI_ISL_463547|2020-04-3 .................... .................... .................... .................... .................... ....................
D 7730 hCoV-19/USA/WA-S1058/2020|EPI_ISL_463548|2020-04-2 .................... .................... .................... .................... .................... ....................
D 7731 hCoV-19/USA/WA-S1059/2020|EPI_ISL_463549|2020-04-2 .................... .................... .................... .................... .................... ....................
D 7732 hCoV-19/USA/WA-S1060/2020|EPI_ISL_463550|2020-04-3 .................... .................... .................... .................... .................... ....................
D 7733 hCoV-19/USA/WA-S1061/2020|EPI_ISL_463551|2020-04-3 .................... .................... .................... .................... .................... ....................
D 7734 hCoV-19/USA/WA-S1062/2020|EPI_ISL_463552|2020-04-3 .................... .................... .................... .................... .................... ....................
D 7735 hCoV-19/USA/WA-S1063/2020|EPI_ISL_463553|2020-04-3 .................... .................... .................... .................... .................... ....................
D 7736 hCoV-19/USA/WA-S1064/2020|EPI_ISL_463554|2020-04-3 .................... .................... .................... .................... .................... ....................
D 7737 hCoV-19/USA/WA-S1065/2020|EPI_ISL_463555|2020-04-2 .................... .................... .................... .................... .................... ....................
D 7738 hCoV-19/USA/WA-S1066/2020|EPI_ISL_463556|2020-04-2 .................... .................... .................... .................... .................... ....................
D 7739 hCoV-19/USA/WA-S1067/2020|EPI_ISL_463557|2020-04-2 .................... .................... .................... .................... .................... ....................
D 7740 hCoV-19/USA/WA-S1068/2020|EPI_ISL_463558|2020-04-2 .................... .................... .................... .................... .................... ....................
D 7741 hCoV-19/USA/WA-S1069/2020|EPI_ISL_463559|2020-04-3 .................... .................... .................... .................... .................... ....................
D 7742 hCoV-19/USA/WA-S1070/2020|EPI_ISL_463560|2020-04-3 .................... .................... .................... .................... .................... ....................
D 7743 hCoV-19/USA/WA-S1071/2020|EPI_ISL_463561|2020-04-3 .................... .................... .................... .................... .................... ....................

D 7744 hCoV-19/USA/WA-S1072/2020|EPI_ISL_463562|2020-04-3 ···················· ···················· ···················· ···················· ···················· ····················
D 7745 hCoV-19/USA/WA-S1073/2020|EPI_ISL_463563|2020-04-3 ···················· ···················· ···················· ···················· ···················· ····················
D 7747 hCoV-19/England/201180001/2020|EPI_ISL_464874|2020 ···················· ···················· ···················· ···················· ···················· ····················
D 7750 hCoV-19/England/201180067/2020|EPI_ISL_464879|2020 ···················· ···················· ···················· ···················· ···················· ····················
D 7751 hCoV-19/England/20118006904/2020|EPI_ISL_464880|20 ···················· ···················· ···················· ···················· ·NNN················ ···················· 1
D 7754 hCoV-19/England/201180317/2020|EPI_ISL_464894|2020 ···················· ···················· ···················· ···················· ···················· ····················
D 7756 hCoV-19/Switzerland/BE-ETHZ-120030/2020|EPI_ISL_46 ···················· ···················· ···················· ···················· ···················· ····················
D 7757 hCoV-19/Switzerland/ZH-ETHZ-120031/2020|EPI_ISL_46 ···················· ···················· ···················· ···················· ···················· ····················
D 7762 hCoV-19/Switzerland/SG-ETHZ-120040/2020|EPI_ISL_46 ···················· ···················· ···················· ···················· ···················· ····················
D 7764 hCoV-19/USA/CA-CZB-1430/2020|EPI_ISL_468400|2020-0 ···················· ···················· ···················· ···················· ···················· ····················
D 7765 hCoV-19/USA/CA-CZB-1431/2020|EPI_ISL_468401|2020-0 ···················· ···················· ···················· ···················· ···················· ····················
D 7766 hCoV-19/USA/CA-CZB-1432/2020|EPI_ISL_468402|2020-0 ···················· ···················· ···················· ···················· ···················· ····················
D 7767 hCoV-19/USA/CA-CZB-1433/2020|EPI_ISL_468403|2020-0 ···················· ···················· ···················· ···················· ···················· ····················
D 7768 hCoV-19/USA/CA-CZB-1434/2020|EPI_ISL_468404|2020-0 ···················· ···················· ···················· ···················· ···················· ····················
D 7769 hCoV-19/USA/CA-CZB-1435/2020|EPI_ISL_468405|2020-0 ···················· ···················· ···················· ···················· ···················· ····················
D 7770 hCoV-19/USA/CA-CZB-1436/2020|EPI_ISL_468406|2020-0 ···················· ···················· ···················· ···················· ···················· ····················
D 7771 hCoV-19/USA/CA-CZB-1437/2020|EPI_ISL_468407|2020-0 ···················· ···················· ···················· ···················· ···················· ····················
D 7772 hCoV-19/USA/CA-CZB-1442/2020|EPI_ISL_468409|2020-0 ···················· ···················· ···················· ···················· ···················· ····················
D 7773 hCoV-19/USA/CA-CZB-1443/2020|EPI_ISL_468410|2020-0 ···················· ···················· ···················· ···················· ···················· ····················
D 7774 hCoV-19/USA/CA-CZB-1445/2020|EPI_ISL_468411|2020-0 ···················· ···················· ···················· ···················· ···················· ····················
D 7775 hCoV-19/USA/CA-CZB-1447/2020|EPI_ISL_468413|2020-0 ···················· ···················· ···················· ···················· ···················· ····················
D 7776 hCoV-19/USA/CA-CZB-1450/2020|EPI_ISL_468414|2020-0 ···················· ···················· ···················· ···················· ···················· ····················
D 7777 hCoV-19/England/BRIS-12759A/2020|EPI_ISL_469548|20 ···················· ···················· ···················· ···················· ···················· ····················
D 7778 hCoV-19/England/BRIS-1270D5/2020|EPI_ISL_469549|20 ···················· ···················· ···················· ···················· ···················· ····················
D 7779 hCoV-19/England/BRIS-127F9C/2020|EPI_ISL_469550|20 ···················· ···················· ···················· ···················· ···················· ····················
D 7780 hCoV-19/England/BRIS-1275E5/2020|EPI_ISL_469551|20 ···················· ···················· ···················· ···················· ···················· ····················
D 7781 hCoV-19/England/BRIS-1277D0/2020|EPI_ISL_469553|20 ···················· ···················· ···················· ···················· ···················· ····················
D 7782 hCoV-19/England/BRIS-127864/2020|EPI_ISL_469556|20 ···················· ···················· ···················· ···················· ···················· ····················
D 7783 hCoV-19/England/BRIS-127B5C/2020|EPI_ISL_469557|20 ···················· ···················· ···················· ···················· ···················· ····················
D 7784 hCoV-19/England/BRIS-126EAF/2020|EPI_ISL_469558|20 ···················· ···················· ···················· ···················· ···················· ····················
D 7785 hCoV-19/England/BRIS-1280C5/2020|EPI_ISL_469559|20 ···················· ···················· ···················· ···················· ···················· ····················
D 7786 hCoV-19/England/BRIS-1274E8/2020|EPI_ISL_469560|20 ···················· ···················· ···················· ···················· ···················· ····················
D 7787 hCoV-19/England/BRIS-128371/2020|EPI_ISL_469566|20 ···················· ···················· ···················· ···················· ···················· ····················
D 7788 hCoV-19/England/BRIS-127846/2020|EPI_ISL_469568|20 ···················· ···················· ···················· ···················· ···················· ····················
D 7789 hCoV-19/England/BRIS-127C68/2020|EPI_ISL_469700|20 ···················· ···················· ···················· ···················· ···················· ····················
D 7790 hCoV-19/England/BRIS-12714B/2020|EPI_ISL_469701|20 ···················· ···················· ···················· ···················· ···················· ····················
D 7791 hCoV-19/England/BRIS-1279AD/2020|EPI_ISL_469704|20 ···················· ···················· ···················· ···················· ···················· ····················
D 7792 hCoV-19/England/BRIS-126F51/2020|EPI_ISL_469705|20 ···················· ···················· ···················· ···················· ···················· ····················
D 7793 hCoV-19/England/BRIS-127F8D/2020|EPI_ISL_469706|20 ···················· ···················· ···················· ···················· ···················· ····················
D 7794 hCoV-19/England/BRIS-12765B/2020|EPI_ISL_469708|20 ···················· ···················· ···················· ···················· ···················· ····················
D 7795 hCoV-19/England/BRIS-127A5F/2020|EPI_ISL_469709|20 ···················· ···················· ···················· ···················· ···················· ····················
D 7796 hCoV-19/England/BRIS-126FBB/2020|EPI_ISL_469710|20 ···················· ···················· ···················· ···················· ···················· ····················
D 7797 hCoV-19/England/BRIS-128098/2020|EPI_ISL_469711|20 ···················· ···················· ···················· ···················· ···················· ····················
D 7798 hCoV-19/England/BRIS-127C2C/2020|EPI_ISL_469712|20 ···················· ···················· ···················· ···················· ···················· ····················
D 7799 hCoV-19/England/BRIS-12706C/2020|EPI_ISL_469713|20 ···················· ···················· ···················· ···················· ···················· ····················
D 7800 hCoV-19/England/BRIS-12804D/2020|EPI_ISL_469714|20 ···················· ···················· ···················· ···················· ···················· ····················
D 7801 hCoV-19/England/BRIS-128450/2020|EPI_ISL_469715|20 ···················· ···················· ···················· ···················· ···················· ····················
D 7802 hCoV-19/England/BRIS-127BD4/2020|EPI_ISL_469720|20 ···················· ···················· ···················· ···················· ···················· ····················
D 7803 hCoV-19/England/BRIS-128405/2020|EPI_ISL_469721|20 ············T······· ···················· ···················· ···················· ···················· ····················
D 7804 hCoV-19/England/BRIS-126EEB/2020|EPI_ISL_469723|20 ···················· ····A··············· ···················· ···················· ···················· ····················
D 7805 hCoV-19/England/BRIS-127A40/2020|EPI_ISL_469724|20 ···················· ···················· ···················· ···················· ···················· ····················
D 7806 hCoV-19/England/BRIS-12704E/2020|EPI_ISL_469725|20 ···················· ···················· ···················· ···················· ···················· ····················
D 7807 hCoV-19/England/BRIS-126FD9/2020|EPI_ISL_469726|20 ···················· ···················· ···················· ···················· ···················· ····················
D 7808 hCoV-19/England/BRIS-128380/2020|EPI_ISL_469727|20 ···················· ···················· ···················· ···················· ···················· ····················
D 7809 hCoV-19/England/BRIS-127F50/2020|EPI_ISL_469728|20 ···················· ·············A······ ···················· ···················· ···················· ····················
D 7810 hCoV-19/England/BRIS-127FC9/2020|EPI_ISL_469730|20 ···················· ···················· ···················· ···················· ···················· ····················
D 7811 hCoV-19/England/BRIS-1285E4/2020|EPI_ISL_469731|20 ···················· ···················· ···················· ···················· ···················· ····················
D 7812 hCoV-19/England/BRIS-127196/2020|EPI_ISL_469733|20 ···················· ···················· ···················· ···················· ···················· ····················
D 7813 hCoV-19/England/BRIS-1271B4/2020|EPI_ISL_469735|20 ···················· ···················· ···················· ···················· ···················· ····················
D 7814 hCoV-19/England/BRIS-127A6E/2020|EPI_ISL_469737|20 ···················· ···················· ···················· ···················· ···················· ····················
D 7815 hCoV-19/England/BRIS-127A9B/2020|EPI_ISL_469738|20 ···················· ····A··············· ···················· ···················· ···················· ····················
D 7816 hCoV-19/England/BRIS-126FF7/2020|EPI_ISL_469739|20 ···················· ···················· ···················· ···················· ···················· ····················
D 7817 hCoV-19/England/BRIS-1281A4/2020|EPI_ISL_469740|20 ···················· ···················· ···················· ···················· ···················· ····················
D 7818 hCoV-19/England/BRIS-126E90/2020|EPI_ISL_469741|20 ···················· ···················· ···················· ···················· ···················· ····················
D 7819 hCoV-19/England/BRIS-126F33/2020|EPI_ISL_469742|20 ···················· ···················· ···················· ···················· ···················· ····················
D 7820 hCoV-19/England/BRIS-1274CA/2020|EPI_ISL_469743|20 ···················· ···················· ···················· ···················· ···················· ····················
D 7821 hCoV-19/England/BRIS-1279E9/2020|EPI_ISL_469744|20 ···················· ···················· ···················· ···················· ···················· ····················
D 7822 hCoV-19/England/BRIS-126F06/2020|EPI_ISL_469745|20 ···················· ····A··············· ···················· ···················· ···················· ····················
D 7823 hCoV-19/England/CAMB-7DE9B/2020|EPI_ISL_470513|202 ···················· ···················· ···················· ···················· ···················· ····················
D 7824 hCoV-19/England/CAMB-801C9/2020|EPI_ISL_470514|202 ···················· ···················· ···················· ···················· ···················· ····················
D 7825 hCoV-19/England/CAMB-7BE9D/2020|EPI_ISL_470515|202 ···················· ···················· ···················· ···················· ···················· ····················
D 7826 hCoV-19/England/CAMB-803D2/2020|EPI_ISL_470516|202 ···················· ···················· ···················· ···················· ···················· ····················
D 7827 hCoV-19/England/CAMB-809EF/2020|EPI_ISL_470519|202 ···················· ···················· ···················· ···················· ···················· ····················
D 7828 hCoV-19/England/CAMB-7BEBB/2020|EPI_ISL_470520|202 ···················· ···················· ···················· ···················· ···················· ····················
D 7829 hCoV-19/England/CAMB-7F5C1/2020|EPI_ISL_470521|202 ···················· ···················· ···················· ···················· ···················· ····················
D 7830 hCoV-19/England/CAMB-805BE/2020|EPI_ISL_470522|202 ···················· ···················· ···················· ···················· ···················· ····················
D 7831 hCoV-19/England/CAMB-805CD/2020|EPI_ISL_470523|202 ···················· ···················· ···················· ···················· ···················· ····················
D 7832 hCoV-19/England/CAMB-7F52B/2020|EPI_ISL_470524|202 ···················· ···················· ···················· ···················· ···················· ····················

| | | | | | | | | |
|---|---|---|---|---|---|---|---|---|
| D | 7833 | hCoV-19/England/CAMB-803C3/2020\|EPI_ISL_470526\|202 | ···················· | ···················· | ···················· | ···················· | ···················· | ···················· |
| D | 7834 | hCoV-19/England/CAMB-80132/2020\|EPI_ISL_470527\|202 | ···················· | ···················· | ···················· | ···················· | ···················· | ···················· |
| D | 7835 | hCoV-19/England/CAMB-80105/2020\|EPI_ISL_470528\|202 | ···················· | ···················· | ···················· | ···················· | ···················· | ···················· |
| D | 7836 | hCoV-19/England/BRIS-123983/2020\|EPI_ISL_470529\|20 | ···················· | ···················· | ···················· | ···················· | ···················· | ···················· |
| D | 7837 | hCoV-19/USA/UT-01878/2020\|EPI_ISL_470700\|2020-04-1 | ···················· | ···················· | ···················· | ···················· | ···················· | ···················· |
| D | 7838 | hCoV-19/USA/UT-01881/2020\|EPI_ISL_470701\|2020-04-1 | ···················· | ···················· | ···················· | ···················· | ···················· | ···················· |
| D | 7839 | hCoV-19/USA/UT-01883/2020\|EPI_ISL_470703\|2020-04-1 | ···················· | ···················· | ···················· | ···················· | ···················· | ···················· |
| D | 7840 | hCoV-19/USA/UT-01884/2020\|EPI_ISL_470704\|2020-04-1 | ···················· | ···················· | ···················· | ···················· | ···················· | ···················· |
| D | 7841 | hCoV-19/USA/UT-01885/2020\|EPI_ISL_470705\|2020-04-1 | ···················· | ···················· | ···················· | ···················· | ···················· | ···················· |
| D | 7842 | hCoV-19/USA/UT-01886/2020\|EPI_ISL_470706\|2020-04-1 | ···················· | ···················· | ···················· | ···················· | ···················· | ···················· |
| D | 7843 | hCoV-19/USA/UT-02136/2020\|EPI_ISL_470708\|2020-04-1 | ···················· | ···················· | ···················· | ···················· | ···················· | ···················· |
| D | 7844 | hCoV-19/USA/UT-02140/2020\|EPI_ISL_470711\|2020-04-1 | ···················· | ···················· | ···················· | ···················· | ···················· | ···················· |
| D | 7845 | hCoV-19/USA/UT-03810/2020\|EPI_ISL_470712\|2020-04-0 | ···················· | ···················· | ···················· | ···················· | ···················· | ···················· |
| D | 7846 | hCoV-19/USA/UT-03857/2020\|EPI_ISL_470716\|2020-04-0 | ···················· | ···················· | ···················· | ···················· | ···················· | ···················· |
| D | 7847 | hCoV-19/USA/UT-03858/2020\|EPI_ISL_470717\|2020-04-0 | ···················· | ···················· | ···················· | ···················· | ···················· | ···················· |
| D | 7848 | hCoV-19/USA/UT-03860/2020\|EPI_ISL_470721\|2020-04-0 | ···················· | ···················· | ···················· | ···················· | ···················· | ···················· |
| D | 7849 | hCoV-19/USA/UT-03867/2020\|EPI_ISL_470724\|2020-04-0 | ···················· | ···················· | ···················· | ···················· | ···················· | ···················· |
| D | 7850 | hCoV-19/USA/UT-03869/2020\|EPI_ISL_470725\|2020-04-0 | ···················· | ···················· | ···················· | ···················· | ···················· | ···················· |
| D | 7851 | hCoV-19/USA/UT-03872/2020\|EPI_ISL_470728\|2020-04-0 | ···················· | ···················· | ···················· | ···················· | ···················· | ···················· |
| D | 7852 | hCoV-19/USA/MI-MDHHS-SC20823/2020\|EPI_ISL_471844\|2 | ···················· | ··C················· | ···················· | ···················· | ···················· | ···················· |
| D | 7853 | hCoV-19/USA/MI-MDHHS-SC20826/2020\|EPI_ISL_471847\|2 | ···················· | ···················· | ···················· | ···················· | ···················· | ···················· |
| D | 7854 | hCoV-19/USA/MI-MDHHS-SC20827/2020\|EPI_ISL_471848\|2 | ···················· | ···················· | ···················· | ···················· | ···················· | ···················· |
| D | 7855 | hCoV-19/USA/MI-MDHHS-SC20829/2020\|EPI_ISL_471850\|2 | ···················· | ···················· | ···················· | ···················· | ···················· | ···················· |
| D | 7856 | hCoV-19/USA/MI-MDHHS-SC20830/2020\|EPI_ISL_471851\|2 | ···················· | ···················· | ···················· | ···················· | ···················· | ···················· |
| D | 7857 | hCoV-19/USA/MI-MDHHS-SC20831/2020\|EPI_ISL_471852\|2 | ···················· | ···················· | ···················· | ···················· | ···················· | ···················· |
| D | 7858 | hCoV-19/USA/MI-MDHHS-SC20832/2020\|EPI_ISL_471853\|2 | ···················· | ···················· | ···················· | ···················· | ···················· | ···················· |
| D | 7859 | hCoV-19/USA/MI-MDHHS-SC20833/2020\|EPI_ISL_471854\|2 | ···················· | ···················· | ···················· | ···················· | ···················· | ···················· |
| D | 7860 | hCoV-19/USA/MI-MDHHS-SC20834/2020\|EPI_ISL_471855\|2 | ···················· | ···················· | ···················· | ···················· | ···················· | ···················· |
| D | 7861 | hCoV-19/USA/MI-MDHHS-SC20835/2020\|EPI_ISL_471856\|2 | ···················· | ···················· | ···················· | ···················· | ···················· | ···················· |
| D | 7862 | hCoV-19/USA/MI-MDHHS-SC20836/2020\|EPI_ISL_471857\|2 | ···················· | ···················· | ···················· | ···················· | ···················· | ···················· |
| D | 7863 | hCoV-19/USA/MI-MDHHS-SC20837/2020\|EPI_ISL_471858\|2 | ···················· | ···················· | ···················· | ···················· | ···················· | ···················· |
| D | 7864 | hCoV-19/USA/MI-MDHHS-SC20838/2020\|EPI_ISL_471859\|2 | ···················· | ···················· | ···················· | ···················· | ···················· | ···················· |
| D | 7865 | hCoV-19/Wales/PHWC-16468F/2020\|EPI_ISL_474212\|2020 | ···················· | ···················· | ···················· | ···················· | ···················· | ···················· |
| D | 7866 | hCoV-19/Wales/PHWC-1646AD/2020\|EPI_ISL_474214\|2020 | ···················· | ···················· | ···················· | ···················· | ···················· | ···················· |
| D | 7867 | hCoV-19/Wales/PHWC-1646BC/2020\|EPI_ISL_474215\|2020 | ···················· | ···················· | ···················· | ···················· | ···················· | ···················· |
| D | 7868 | hCoV-19/Wales/PHWC-1646CB/2020\|EPI_ISL_474216\|2020 | ···················· | ···················· | ···················· | ···················· | ···················· | ···················· |
| D | 7869 | hCoV-19/Wales/PHWC-1646DA/2020\|EPI_ISL_474217\|2020 | ···················· | ···················· | ···················· | ···················· | ···················· | ···················· |
| D | 7870 | hCoV-19/Wales/PHWC-164704/2020\|EPI_ISL_474220\|2020 | ···················· | ···················· | ···················· | ···················· | ···················· | ···················· |
| D | 7871 | hCoV-19/Wales/PHWC-24051/2020\|EPI_ISL_474225\|2020- | ···················· | ···················· | ···················· | ···················· | ···················· | ···················· |
| D | 7872 | hCoV-19/Wales/PHWC-29AF4/2020\|EPI_ISL_474239\|2020- | ···················· | ···················· | ···················· | ···················· | ···················· | ···················· |
| D | 7873 | hCoV-19/Wales/PHWC-2AE06/2020\|EPI_ISL_474259\|2020- | ···················· | ···················· | ···················· | ···················· | ···················· | ···················· |
| D | 7874 | hCoV-19/Wales/PHWC-2AFA9/2020\|EPI_ISL_474260\|2020- | ···················· | ···················· | ···················· | ···················· | ···················· | ···················· |
| D | 7875 | hCoV-19/Wales/PHWC-2BFD5/2020\|EPI_ISL_474265\|2020- | ···················· | ···················· | ···················· | ···················· | ···················· | ···················· |
| D | 7876 | hCoV-19/Wales/PHWC-2C0E0/2020\|EPI_ISL_474275\|2020- | ···················· | ···················· | ···················· | ···················· | ···················· | ···················· |
| D | 7877 | hCoV-19/Wales/PHWC-2C1FC/2020\|EPI_ISL_474291\|2020- | ···················· | ···················· | ···················· | ···················· | ···················· | ···················· |
| D | 7878 | hCoV-19/Wales/PHWC-2C217/2020\|EPI_ISL_474293\|2020- | ···················· | ···················· | ···················· | ···················· | ···················· | ···················· |
| D | 7879 | hCoV-19/Wales/PHWC-2C244/2020\|EPI_ISL_474296\|2020- | ···················· | ···················· | ···················· | ···················· | ···················· | ···················· |
| D | 7880 | hCoV-19/Sweden/20-51351/2020\|EPI_ISL_475543\|2020-0 | ···················· | ···················· | ···················· | ···················· | ···················· | ···················· |
| D | 7881 | hCoV-19/Sweden/20-04736/2020\|EPI_ISL_475544\|2020-0 | ···················· | ···················· | ···················· | ···················· | ···················· | ···················· |
| D | 7882 | hCoV-19/Sweden/20-04850/2020\|EPI_ISL_475545\|2020-0 | ···················· | ···················· | ···················· | ···················· | ···················· | ···················· |
| D | 7883 | hCoV-19/Sweden/20-04851/2020\|EPI_ISL_475546\|2020-0 | ···················· | ···················· | ···················· | ···················· | ···················· | ···················· |
| D | 7884 | hCoV-19/Sweden/20-04852/2020\|EPI_ISL_475547\|2020-0 | ···················· | ···················· | ···················· | ···················· | ···················· | ···················· |
| D | 7885 | hCoV-19/Sweden/20-08041/2020\|EPI_ISL_475548\|2020-0 | ···················· | ···················· | ···················· | ···················· | ···················· | ···················· |
| D | 7886 | hCoV-19/Sweden/20-08046/2020\|EPI_ISL_475549\|2020-0 | ···················· | ···················· | ···················· | ···················· | ···················· | ···················· |
| D | 7887 | hCoV-19/Sweden/20-08056/2020\|EPI_ISL_475550\|2020-0 | ···················· | ···················· | ···················· | ···················· | ···················· | ···················· |
| D | 7888 | hCoV-19/Sweden/20-08067/2020\|EPI_ISL_475551\|2020-0 | ···················· | ···················· | ···················· | ···················· | ···················· | ···················· |
| D | 7889 | hCoV-19/Sweden/20-08074/2020\|EPI_ISL_475552\|2020-0 | ···················· | ···················· | ···················· | ···················· | ···················· | ···················· |
| D | 7890 | hCoV-19/Sweden/20-08133/2020\|EPI_ISL_475553\|2020-0 | ···················· | ···················· | ···················· | ···················· | ···················· | ···················· |
| D | 7891 | hCoV-19/Sweden/20-08136/2020\|EPI_ISL_475554\|2020-0 | ···················· | ···················· | ···················· | ···················· | ···················· | ···················· |
| D | 7892 | hCoV-19/Sweden/20-08160/2020\|EPI_ISL_475555\|2020-0 | ···················· | ···················· | ···················· | ···················· | ···················· | ···················· |
| D | 7893 | hCoV-19/Sweden/20-08639/2020\|EPI_ISL_475556\|2020-0 | ···················· | ···················· | ···················· | ···················· | ···················· | ···················· |
| D | 7894 | hCoV-19/Sweden/20-08840/2020\|EPI_ISL_475557\|2020-0 | ···················· | ···················· | ···················· | ···················· | ···················· | ···················· |
| D | 7895 | hCoV-19/Sweden/20-50204/2020\|EPI_ISL_475558\|2020-0 | ···················· | ···················· | ···················· | ···················· | ···················· | ···················· |
| D | 7896 | hCoV-19/Sweden/20-50237/2020\|EPI_ISL_475560\|2020-0 | ···················· | ···················· | ···················· | ···················· | ···················· | ···················· |
| D | 7897 | hCoV-19/Sweden/20-50243/2020\|EPI_ISL_475561\|2020-0 | ···················· | ···················· | ···················· | ···················· | ···················· | ···················· |
| D | 7898 | hCoV-19/Sweden/20-05983/2020\|EPI_ISL_475562\|2020-0 | ···················· | ···················· | ···················· | ···················· | ···················· | ···················· |
| D | 7899 | hCoV-19/Sweden/20-05984/2020\|EPI_ISL_475563\|2020-0 | ···················· | ···················· | ···················· | ···················· | ···················· | ···················· |
| D | 7900 | hCoV-19/Sweden/20-06523/2020\|EPI_ISL_475564\|2020-0 | ···················· | ···················· | ···················· | ···················· | ···················· | ···················· |
| D | 7901 | hCoV-19/Sweden/20-19258/2020\|EPI_ISL_475567\|2020-0 | ···················· | ···················· | ···················· | ···················· | ···················· | ···················· |
| D | 7902 | hCoV-19/Sweden/20-51816/2020\|EPI_ISL_475568\|2020-0 | ···················· | ···················· | ···················· | ···················· | ···················· | ···················· |
| D | 7903 | hCoV-19/Sweden/20-51988/2020\|EPI_ISL_475569\|2020-0 | ···················· | ···················· | ···················· | ···················· | ···················· | ···················· |
| D | 7904 | hCoV-19/USA/CA-CSMC112/2020\|EPI_ISL_475588\|2020-04 | ···················· | ···················· | ···················· | ···················· | ···················· | ···················· |
| D | 7905 | hCoV-19/USA/CA-CSMC114/2020\|EPI_ISL_475590\|2020-04 | ···················· | ···················· | ···················· | ···················· | ···················· | ···················· |
| D | 7906 | hCoV-19/USA/CA-CSMC117/2020\|EPI_ISL_475593\|2020-04 | ···················· | ···················· | ···················· | ···················· | ···················· | ···················· |
| D | 7907 | hCoV-19/USA/CA-CSMC118/2020\|EPI_ISL_475594\|2020-04 | ···················· | ···················· | ···················· | ···················· | ···················· | ···················· |
| D | 7908 | hCoV-19/USA/CA-CSMC119/2020\|EPI_ISL_475595\|2020-04 | ···················· | ···················· | ···················· | ···················· | ···················· | ···················· |
| D | 7909 | hCoV-19/USA/CA-CSMC12/2020\|EPI_ISL_475596\|2020-03- | ···················· | ···················· | ···················· | ···················· | ···················· | ···················· |
| D | 7910 | hCoV-19/USA/CA-CSMC121/2020\|EPI_ISL_475598\|2020-04 | ···················· | ···················· | ···················· | ···················· | ···················· | ···················· |

| | | | | | | | |
|---|---|---|---|---|---|---|---|
| D | 7911 hCoV-19/USA/CA-CSMC122/2020\|EPI_ISL_475599\|2020-04 | ···················· | ···················· | ···················· | ···················· | ···················· | ···················· |
| D | 7912 hCoV-19/India/GJ-GBRC233a/2020\|EPI_ISL_476874\|2020 | ···················· | ···················· | ···················· | ···················· | ···················· | ···················· |
| D | 7913 hCoV-19/India/GJ-GBRC233b/2020\|EPI_ISL_476875\|2020 | ···················· | ···················· | ···················· | ···················· | ···················· | ···················· |
| D | 7914 hCoV-19/India/GJ-GBRC235a/2020\|EPI_ISL_476876\|2020 | ···················· | ···················· | ···················· | ···················· | ···················· | ··A················· |
| D | 7915 hCoV-19/India/GJ-GBRC235b/2020\|EPI_ISL_476877\|2020 | ···················· | ·A·················· | ···················· | ···················· | ···················· | ···················· |
| D | 7916 hCoV-19/India/GJ-GBRC236/2020\|EPI_ISL_476878\|2020- | ···················· | ···················· | ···················· | ···················· | ···················· | ···················· |
| D | 7917 hCoV-19/India/GJ-GBRC237a/2020\|EPI_ISL_476879\|2020 | ···················· | ···················· | ···················· | ···················· | ···················· | ···················· |
| D | 7918 hCoV-19/India/GJ-GBRC237b/2020\|EPI_ISL_476880\|2020 | ···················· | ···················· | ···················· | ···················· | ···················· | ···················· |
| D | 7919 hCoV-19/India/GJ-GBRC238a/2020\|EPI_ISL_476881\|2020 | ···················· | ···················· | ···················· | ···················· | ···················· | ···················· |
| D | 7920 hCoV-19/India/GJ-GBRC238b/2020\|EPI_ISL_476882\|2020 | ···················· | ···················· | ···················· | ···················· | ···················· | ··A················· |
| D | 7921 hCoV-19/India/MP-DRDE 3510/2020\|EPI_ISL_476883\|202 | ···················· | ···················· | ···················· | ···················· | ···················· | ···················· |
| D | 7922 hCoV-19/India/MP-DRDE 3926/2020\|EPI_ISL_476884\|202 | ···················· | ···················· | ···················· | ···················· | ···················· | ···················· |
| D | 7923 hCoV-19/India/MP-DRDE 4078/2020\|EPI_ISL_476885\|202 | ···················· | ···················· | ···················· | ···················· | ·········M·········· | ···················· |
| D | 7924 hCoV-19/India/MP-DRDE 4083/2020\|EPI_ISL_476886\|202 | ···················· | ···················· | ···················· | ···················· | ···················· | ···················· |
| D | 7925 hCoV-19/India/MP-DRDE-4093/2020\|EPI_ISL_476887\|202 | ···················· | ···················· | ···················· | ···················· | ···················· | ···················· |
| D | 7926 hCoV-19/India/MP-DRDE-3983/2020\|EPI_ISL_476888\|202 | ···················· | ···················· | ···················· | ···················· | ···················· | ···················· |
| D | 7927 hCoV-19/India/MP-DRDE-3987/2020\|EPI_ISL_476889\|202 | ···················· | ·R·················· | ···················· | ···················· | ···················· | ··R················· |
| D | 7928 hCoV-19/India/MP-DRDE-4022/2020\|EPI_ISL_476890\|202 | ···················· | ···················· | ···················· | ···················· | ······S··W········· | ···················· |
| D | 7929 hCoV-19/India/MP-DRDE-4096/2020\|EPI_ISL_476891\|202 | ···················· | ···················· | ···················· | ···················· | ···················· | ···················· |
| D | 7930 hCoV-19/India/MP-DRDE-4105/2020\|EPI_ISL_476892\|202 | ···················· | ···················· | ···················· | ···················· | ···················· | ···················· |
| D | 7931 hCoV-19/India/MP-DRDE-4111/2020\|EPI_ISL_476893\|202 | ···················· | ···················· | ···················· | ···················· | ···················· | ···················· |
| D | 7932 hCoV-19/India/MP-DRDE-4195/2020\|EPI_ISL_476894\|202 | ···················· | ···················· | ···················· | ···················· | ···················· | ···················· |
| D | 7933 hCoV-19/India/MP-DRDE 4661/2020\|EPI_ISL_476895\|202 | ···················· | ···················· | ···················· | ···················· | ···················· | ···················· |
| D | 7934 hCoV-19/India/MP-DRDE-4624/2020\|EPI_ISL_476896\|202 | ···················· | ···················· | ···················· | ···················· | ···················· | ···················· |
| D | 7935 hCoV-19/USA/AK-PHL141/2020\|EPI_ISL_476898\|2020-03- | ···················· | ···················· | ···················· | ···················· | ···················· | ···················· |
| D | 7936 hCoV-19/USA/AK-PHL142/2020\|EPI_ISL_476899\|2020-03- | ···················· | ···················· | ···················· | ···················· | ···················· | ···················· |
| D | 7937 hCoV-19/Belgium/VME-0327346/2020\|EPI_ISL_462200\|20 | ···················· | ···················· | ···················· | ···················· | ···················· | ···················· |
| D | 7938 hCoV-19/Belgium/BN-0330347/2020\|EPI_ISL_462201\|202 | ···················· | ···················· | ···················· | ···················· | ···················· | ···················· |
| D | 7939 hCoV-19/Belgium/FCA-0327348/2020\|EPI_ISL_462202\|20 | ···················· | ···················· | ···················· | ···················· | ···················· | ···················· |
| D | 7940 hCoV-19/Belgium/GGO-0327349/2020\|EPI_ISL_462203\|20 | ···················· | ···················· | ···················· | ···················· | ···················· | ···················· |
| D | 7941 hCoV-19/Belgium/DC-0326350/2020\|EPI_ISL_462204\|202 | ···················· | ···················· | ···················· | ···················· | ···················· | ···················· |
| D | 7942 hCoV-19/Belgium/DNM-0330351/2020\|EPI_ISL_462205\|20 | ···················· | ···················· | ···················· | ···················· | ···················· | ···················· |
| D | 7943 hCoV-19/Belgium/VEH-0329352/2020\|EPI_ISL_462206\|20 | ···················· | ···················· | ···················· | ···················· | ···················· | ···················· |
| D | 7944 hCoV-19/Belgium/DV-0327353/2020\|EPI_ISL_462207\|202 | ···················· | ···················· | ···················· | ···················· | ···················· | ···················· |
| D | 7945 hCoV-19/Belgium/GJM-0328354/2020\|EPI_ISL_462208\|20 | ···················· | ···················· | ···················· | ···················· | ···················· | ···················· |
| D | 7946 hCoV-19/Belgium/DC-0322355/2020\|EPI_ISL_462209\|202 | ···················· | ···················· | ···················· | ···················· | ···················· | ···················· |
| D | 7947 hCoV-19/Belgium/BWM-0311356/2020\|EPI_ISL_462210\|20 | ···················· | ···················· | ···················· | ···················· | ···················· | ···················· |
| D | 7948 hCoV-19/Belgium/VPP-0331379/2020\|EPI_ISL_462233\|20 | ···················· | ···················· | ···················· | ···················· | ···················· | ···················· |
| D | 7949 hCoV-19/Belgium/SC-0325380/2020\|EPI_ISL_462234\|202 | ···················· | ···················· | ···················· | ···················· | ···················· | ···················· |
| D | 7950 hCoV-19/Belgium/DEJ-0403381/2020\|EPI_ISL_462235\|20 | ···················· | ···················· | ···················· | ···················· | ···················· | ···················· |
| D | 7951 hCoV-19/Belgium/LA-0403382/2020\|EPI_ISL_462236\|202 | ···················· | ···················· | ···················· | ···················· | ···················· | ···················· |
| D | 7952 hCoV-19/Belgium/MG-0402383/2020\|EPI_ISL_462237\|202 | ···················· | ···················· | ···················· | ···················· | ···················· | ···················· |
| D | 7953 hCoV-19/Belgium/LMA-0330384/2020\|EPI_ISL_462238\|20 | ···················· | ···················· | ···················· | ···················· | ···················· | ···················· |
| D | 7954 hCoV-19/Belgium/DLM-0330385/2020\|EPI_ISL_462239\|20 | ···················· | ···················· | ···················· | ···················· | ···················· | ···················· |
| D | 7955 hCoV-19/Belgium/RMO-0329386/2020\|EPI_ISL_462240\|20 | ···················· | ···················· | ···················· | ···················· | ···················· | ···················· |
| D | 7956 hCoV-19/Belgium/VDA-0330387/2020\|EPI_ISL_462241\|20 | ···················· | ···················· | ···················· | ···················· | ···················· | ···················· |
| D | 7957 hCoV-19/Belgium/VSPE-0328388/2020\|EPI_ISL_462242\|2 | ···················· | ···················· | ···················· | ···················· | ···················· | ···················· |
| D | 7958 hCoV-19/Belgium/LHL-0329389/2020\|EPI_ISL_462243\|20 | ···················· | ···················· | ···················· | ···················· | ···················· | ···················· |
| D | 7959 hCoV-19/Belgium/HLE-0327390/2020\|EPI_ISL_462244\|20 | ···················· | ···················· | ···················· | ···················· | ···················· | ···················· |
| D | 7960 hCoV-19/Belgium/TMG-0326391/2020\|EPI_ISL_462245\|20 | ···················· | ···················· | ···················· | ···················· | ···················· | ···················· |
| D | 7961 hCoV-19/Belgium/HAJ-0328392/2020\|EPI_ISL_462246\|20 | ···················· | ···················· | ···················· | ···················· | ···················· | ···················· |
| D | 7962 hCoV-19/Belgium/RIM-0407393/2020\|EPI_ISL_462247\|20 | ···················· | ···················· | ···················· | ···················· | ···················· | ···················· |
| D | 7963 hCoV-19/Belgium/SB-0407394/2020\|EPI_ISL_462248\|202 | ···················· | ···················· | ···················· | ···················· | ···················· | ···················· |
| D | 7964 hCoV-19/Belgium/GM-0407395/2020\|EPI_ISL_462249\|202 | ···················· | ···················· | ···················· | ···················· | ···················· | ···················· |
| D | 7965 hCoV-19/Belgium/JFA-0407396/2020\|EPI_ISL_462250\|20 | ···················· | ···················· | ···················· | ···················· | ···················· | ···················· |
| D | 7966 hCoV-19/Belgium/SP-0407397/2020\|EPI_ISL_462251\|202 | ···················· | ···················· | ···················· | ···················· | ···················· | ···················· |
| D | 7967 hCoV-19/Belgium/DPR-0406398/2020\|EPI_ISL_462252\|20 | ···················· | ···················· | ···················· | ···················· | ···················· | ···················· |
| D | 7968 hCoV-19/Belgium/WCV-0408399/2020\|EPI_ISL_462253\|20 | ···················· | ···················· | ···················· | ···················· | ···················· | ···················· |
| D | 7969 hCoV-19/Belgium/VNM-0406400/2020\|EPI_ISL_462254\|20 | ···················· | ···················· | ···················· | ···················· | ···················· | ···················· |
| D | 7970 hCoV-19/Belgium/REGA-0312401/2020\|EPI_ISL_462255\|2 | ···················· | ···················· | ···················· | ···················· | ···················· | ···················· |
| D | 7971 hCoV-19/Belgium/REGA-0325402/2020\|EPI_ISL_462256\|2 | ···················· | ···················· | ···················· | ···················· | ···················· | ···················· |
| D | 7972 hCoV-19/Belgium/EM-0407403/2020\|EPI_ISL_462257\|202 | ···················· | ···················· | ···················· | ···················· | ···················· | ···················· |
| D | 7973 hCoV-19/Belgium/AO-04074040/2020\|EPI_ISL_462258\|20 | ···················· | ···················· | ···················· | ···················· | ···················· | ···················· |
| D | 7974 hCoV-19/Belgium/PY-0407405/2020\|EPI_ISL_462259\|202 | ···················· | ···················· | ···················· | ···················· | ···················· | ···················· |
| D | 7975 hCoV-19/Belgium/BJA-0409406/2020\|EPI_ISL_462260\|20 | ···················· | ···················· | ···················· | ···················· | ···················· | ···················· |
| D | 7976 hCoV-19/Belgium/DJ-0409407/2020\|EPI_ISL_462261\|202 | ···················· | ···················· | ···················· | ···················· | ···················· | ···················· |
| D | 7977 hCoV-19/Belgium/DMT-0411408/2020\|EPI_ISL_462262\|20 | ···················· | ···················· | ···················· | ···················· | ···················· | ···················· |
| D | 7978 hCoV-19/Belgium/DJ-0323409/2020\|EPI_ISL_462263\|202 | ···················· | ···················· | ···················· | ···················· | ···················· | ···················· |
| D | 7979 hCoV-19/Belgium/KWA-0323410/2020\|EPI_ISL_462264\|20 | ···················· | ···················· | ···················· | ···················· | ···················· | ···················· |
| D | 7980 hCoV-19/Belgium/MT-0322411/2020\|EPI_ISL_462265\|202 | ···················· | ···················· | ···················· | ···················· | ···················· | ···················· |
| D | 7981 hCoV-19/Belgium/AA-0408412/2020\|EPI_ISL_462266\|202 | ···················· | ···················· | ···················· | ···················· | ···················· | ···················· |
| D | 7982 hCoV-19/Belgium/PS-0408413/2020\|EPI_ISL_462267\|202 | ···················· | ···················· | ···················· | ···················· | ···················· | ···················· |
| D | 7983 hCoV-19/Belgium/FM-0408414/2020\|EPI_ISL_462268\|202 | ···················· | ···················· | ···················· | ···················· | ···················· | ···················· |
| D | 7984 hCoV-19/Belgium/DGL-0409415/2020\|EPI_ISL_462269\|20 | ···················· | ···················· | ···················· | ···················· | ···················· | ···················· |
| D | 7985 hCoV-19/Belgium/VAM-0506416/2020\|EPI_ISL_462270\|20 | ···················· | ···················· | ···················· | ···················· | ···················· | ···················· |
| D | 7986 hCoV-19/Belgium/HNF-0507417/2020\|EPI_ISL_462271\|20 | ···················· | ···················· | ···················· | ···················· | ···················· | ···················· |
| D | 7987 hCoV-19/Belgium/VLS-0507418/2020\|EPI_ISL_462272\|20 | ···················· | ···················· | ···················· | ···················· | ···················· | ···················· |
| D | 7988 hCoV-19/Belgium/RJ-0507419/2020\|EPI_ISL_462273\|202 | ···················· | ···················· | ···················· | ···················· | ···················· | ···················· |

| | | | | | | | | | |
|---|---|---|---|---|---|---|---|---|---|
| D | 7989 | hCoV-19/Belgium/SG-0507420/2020\|EPI_ISL_462274\|202 | ···················· | ···················· | ···················· | ···················· | ···················· | ···················· | |
| D | 7990 | hCoV-19/Belgium/HI-0507421/2020\|EPI_ISL_462275\|202 | ···················· | ···················· | ···················· | ···················· | ···················· | ···················· | |
| D | 7991 | hCoV-19/Singapore/158/2020\|EPI_ISL_462276\|2020-04- | ···················· | ···················· | ···················· | ···················· | ···················· | ···················· | |
| D | 7992 | hCoV-19/USA/WA-S1030/2020\|EPI_ISL_463520\|2020-05-0 | ···················· | ···················· | ···················· | ···················· | ···················· | ···················· | |
| D | 7993 | hCoV-19/USA/WA-S1031/2020\|EPI_ISL_463521\|2020-05-0 | ···················· | ···················· | ···················· | ···················· | ···················· | ···················· | |
| D | 7994 | hCoV-19/USA/WA-S1032/2020\|EPI_ISL_463522\|2020-05-0 | ···················· | ···················· | ···················· | ···················· | ···················· | ···················· | |
| D | 7995 | hCoV-19/USA/WA-S1033/2020\|EPI_ISL_463523\|2020-05-0 | ···················· | ···················· | ···················· | ···················· | ···················· | ···················· | |
| D | 7996 | hCoV-19/USA/WA-S1034/2020\|EPI_ISL_463524\|2020-05-0 | ···················· | ···················· | ···················· | ···················· | ···················· | ···················· | |
| D | 7997 | hCoV-19/USA/WA-S1035/2020\|EPI_ISL_463525\|2020-05-0 | ···················· | ···················· | ···················· | ···················· | ···················· | ···················· | |
| D | 7998 | hCoV-19/USA/WA-S1036/2020\|EPI_ISL_463526\|2020-05-0 | ···················· | ···················· | ···················· | ···················· | ···················· | ···················· | |
| D | 7999 | hCoV-19/USA/WA-S1037/2020\|EPI_ISL_463527\|2020-05-0 | ···················· | ···················· | ···················· | ···················· | ···················· | ···················· | |
| D | 8000 | hCoV-19/USA/WA-S1038/2020\|EPI_ISL_463528\|2020-05-0 | ···················· | ···················· | ···················· | ···················· | ···················· | ···················· | |
| D | 8001 | hCoV-19/USA/WA-S1039/2020\|EPI_ISL_463529\|2020-05-0 | ···················· | ···················· | ···················· | ···················· | ···················· | ···················· | |
| D | 8002 | hCoV-19/USA/WA-S1040/2020\|EPI_ISL_463530\|2020-05-0 | ···················· | ···················· | ···················· | ···················· | ···················· | ···················· | |
| D | 8003 | hCoV-19/USA/WA-S1041/2020\|EPI_ISL_463531\|2020-05-0 | ···················· | ···················· | ···················· | ···················· | ···················· | ···················· | |
| D | 8004 | hCoV-19/USA/WA-S1042/2020\|EPI_ISL_463532\|2020-05-0 | ···················· | ···················· | ···················· | ···················· | ···················· | ···················· | |
| D | 8005 | hCoV-19/USA/WA-S1043/2020\|EPI_ISL_463533\|2020-05-0 | ···················· | ···················· | ···················· | ···················· | ···················· | ···················· | |
| D | 8006 | hCoV-19/USA/WA-S1044/2020\|EPI_ISL_463534\|2020-05-0 | ···················· | ···················· | ···················· | ···················· | ···················· | ···················· | |
| D | 8007 | hCoV-19/USA/WA-S1045/2020\|EPI_ISL_463535\|2020-05-0 | ···················· | ···················· | ···················· | ···················· | ···················· | ···················· | |
| D | 8008 | hCoV-19/USA/WA-S1046/2020\|EPI_ISL_463536\|2020-05-0 | ···················· | ···················· | ···················· | ···················· | ···················· | ···················· | |
| D | 8009 | hCoV-19/USA/WA-S1048/2020\|EPI_ISL_463538\|2020-05-0 | ···················· | ·A·················· | ···················· | ···················· | ···················· | ···················· | |
| D | 8010 | hCoV-19/USA/WA-S1049/2020\|EPI_ISL_463539\|2020-05-0 | ···················· | ···················· | ···················· | ···················· | ···················· | ···················· | |
| D | 8011 | hCoV-19/USA/WA-S1050/2020\|EPI_ISL_463540\|2020-04-3 | ···················· | ···················· | ···················· | ···················· | ···················· | ···················· | |
| D | 8012 | hCoV-19/USA/WA-S1051/2020\|EPI_ISL_463541\|2020-04-3 | ···················· | ···················· | ···················· | ···················· | ···················· | ···················· | |
| D | 8013 | hCoV-19/USA/WA-S1074/2020\|EPI_ISL_463564\|2020-04-3 | ···················· | ···················· | ···················· | ···················· | ···················· | ···················· | |
| D | 8014 | hCoV-19/USA/WA-S1075/2020\|EPI_ISL_463565\|2020-04-3 | ···················· | ···················· | ···················· | ···················· | ···················· | ···················· | |
| D | 8015 | hCoV-19/USA/WA-S1076/2020\|EPI_ISL_463566\|2020-04-3 | ···················· | ···················· | ···················· | ···················· | ···················· | ···················· | |
| D | 8016 | hCoV-19/USA/WA-S1077/2020\|EPI_ISL_463567\|2020-04-2 | ···················· | ···················· | ···················· | ···················· | ···················· | ···················· | |
| D | 8017 | hCoV-19/USA/WA-S1078/2020\|EPI_ISL_463568\|2020-04-3 | ···················· | ···················· | ···················· | ···················· | ···················· | ···················· | |
| D | 8018 | hCoV-19/USA/WA-S1079/2020\|EPI_ISL_463569\|2020-04-2 | ···················· | ···················· | ···················· | ···················· | ···················· | ···················· | |
| D | 8019 | hCoV-19/USA/WA-S1080/2020\|EPI_ISL_463570\|2020-04-2 | ···················· | ···················· | ···················· | ···················· | ···················· | ···················· | |
| D | 8020 | hCoV-19/USA/WA-S1081/2020\|EPI_ISL_463571\|2020-04-2 | ···················· | ···················· | ···················· | ···················· | ···················· | ···················· | |
| D | 8021 | hCoV-19/USA/WA-S1082/2020\|EPI_ISL_463572\|2020-04-2 | ···················· | ···················· | ···················· | ···················· | ···················· | ···················· | |
| D | 8022 | hCoV-19/USA/WA-S1083/2020\|EPI_ISL_463573\|2020-04-2 | ···················· | ···················· | ···················· | ···················· | ···················· | ···················· | |
| D | 8023 | hCoV-19/USA/WA-S1084/2020\|EPI_ISL_463574\|2020-04-2 | ···················· | ···················· | ···················· | ···················· | ···················· | ···················· | |
| D | 8024 | hCoV-19/USA/WA-S1085/2020\|EPI_ISL_463575\|2020-04-2 | ···················· | ···················· | ···················· | ···················· | ···················· | ···················· | |
| D | 8025 | hCoV-19/USA/WA-S1086/2020\|EPI_ISL_463576\|2020-04-2 | ···················· | ···················· | ···················· | ···················· | ···················· | ···················· | |
| D | 8026 | hCoV-19/USA/WA-S1087/2020\|EPI_ISL_463577\|2020-04-2 | ···················· | ···················· | ···················· | ···················· | ···················· | ···················· | |
| D | 8027 | hCoV-19/USA/WA-S1089/2020\|EPI_ISL_463579\|2020-04-2 | ···················· | ···················· | ···················· | ···················· | ···················· | ···················· | |
| D | 8028 | hCoV-19/USA/WA-S1090/2020\|EPI_ISL_463580\|2020-04-2 | ···················· | ···················· | ···················· | ···················· | ···················· | ···················· | |
| D | 8029 | hCoV-19/USA/WA-S1091/2020\|EPI_ISL_463581\|2020-04-2 | ···················· | ···················· | ···················· | ···················· | ···················· | ···················· | |
| D | 8030 | hCoV-19/USA/WA-S1092/2020\|EPI_ISL_463582\|2020-04-2 | ···················· | ···················· | ···················· | ···················· | ···················· | ···················· | |
| D | 8031 | hCoV-19/USA/WA-S1093/2020\|EPI_ISL_463583\|2020-05-0 | ···················· | ···················· | ···················· | ···················· | ···················· | ···················· | |
| D | 8032 | hCoV-19/USA/WA-S1094/2020\|EPI_ISL_463584\|2020-05-0 | ···················· | ···················· | ···················· | ···················· | ···················· | ···················· | |
| D | 8033 | hCoV-19/USA/WA-S1095/2020\|EPI_ISL_463585\|2020-05-0 | ···················· | ···················· | ···················· | ···················· | ···················· | ···················· | |
| D | 8034 | hCoV-19/USA/WA-S1096/2020\|EPI_ISL_463586\|2020-05-0 | ···················· | ···················· | ···················· | ···················· | ···················· | ···················· | |
| D | 8035 | hCoV-19/USA/WA-S1097/2020\|EPI_ISL_463587\|2020-05-0 | ···················· | ···················· | ···················· | ···················· | ···················· | ···················· | |
| D | 8036 | hCoV-19/USA/WA-S1098/2020\|EPI_ISL_463588\|2020-05-0 | ···················· | ···················· | ···················· | ···················· | ···················· | ···················· | |
| D | 8037 | hCoV-19/USA/WA-S1099/2020\|EPI_ISL_463589\|2020-05-0 | ···················· | ···················· | ···················· | ···················· | ···················· | ···················· | |
| D | 8038 | hCoV-19/USA/WA-S1100/2020\|EPI_ISL_463590\|2020-05-0 | ···················· | ···················· | ···················· | ···················· | ···················· | ···················· | |
| D | 8039 | hCoV-19/USA/WA-S1101/2020\|EPI_ISL_463591\|2020-05-0 | ···················· | ···················· | ···················· | ···················· | ···················· | ···················· | |
| D | 8040 | hCoV-19/USA/WA-S1102/2020\|EPI_ISL_463592\|2020-05-0 | ···················· | ···················· | ···················· | ···················· | ···················· | ···················· | |
| D | 8041 | hCoV-19/USA/WA-S1103/2020\|EPI_ISL_463593\|2020-05-0 | ···················· | ···················· | ···················· | ···················· | ···················· | ···················· | |
| D | 8042 | hCoV-19/USA/WA-S1104/2020\|EPI_ISL_463594\|2020-05-0 | ···················· | ···················· | ···················· | ···················· | ···················· | ···················· | |
| D | 8043 | hCoV-19/USA/WA-S1105/2020\|EPI_ISL_463595\|2020-05-0 | ···················· | ···················· | ···················· | ···················· | ···················· | ···················· | |
| D | 8044 | hCoV-19/USA/WA-S1106/2020\|EPI_ISL_463596\|2020-05-0 | ···················· | ···················· | ···················· | ···················· | ···················· | ···················· | |
| D | 8045 | hCoV-19/USA/WA-S1107/2020\|EPI_ISL_463597\|2020-05-0 | ···················· | ···················· | ···················· | ···················· | ···················· | ···················· | |
| D | 8046 | hCoV-19/USA/WA-S1108/2020\|EPI_ISL_463598\|2020-05-0 | ···················· | ···················· | ···················· | ···················· | ···················· | ···················· | |
| D | 8047 | hCoV-19/USA/WA-S1109/2020\|EPI_ISL_463599\|2020-05-0 | ···················· | ···················· | ···················· | ···················· | ···················· | ···················· | |
| D | 8048 | hCoV-19/England/201160647/2020\|EPI_ISL_464800\|2020 | ···················· | ···················· | ···················· | ···················· | ···················· | ···················· | |
| D | 8049 | hCoV-19/England/201160658/2020\|EPI_ISL_464801\|2020 | ···················· | ···················· | ···················· | ···················· | ···················· | ···················· | |
| D | 8050 | hCoV-19/England/201160756/2020\|EPI_ISL_464803\|2020 | ···················· | ···················· | ···················· | ···················· | ···················· | ···················· | |
| D | 8051 | hCoV-19/England/201160910/2020\|EPI_ISL_464806\|2020 | ···················· | ···················· | ···················· | ···················· | ···················· | ·NNN················ | 1 |
| D | 8052 | hCoV-19/England/201160912/2020\|EPI_ISL_464807\|2020 | ···················· | ···················· | ···················· | ···················· | ···················· | ···A················ | |
| D | 8053 | hCoV-19/England/201160953/2020\|EPI_ISL_464808\|2020 | ···················· | ···················· | ···················· | ···················· | ···················· | ···················· | |
| D | 8054 | hCoV-19/England/201161525/2020\|EPI_ISL_464852\|2020 | ···················· | ···················· | ···················· | ···················· | ···················· | ···················· | |
| D | 8058 | hCoV-19/England/201161610/2020\|EPI_ISL_464862\|2020 | ···················· | ···················· | ···················· | ···················· | ··N················· | ·N·N················ | |
| D | 8059 | hCoV-19/England/201161611/2020\|EPI_ISL_464863\|2020 | ···················· | ···················· | ···················· | ···················· | ···················· | ···················· | |
| D | 8060 | hCoV-19/England/201161632/2020\|EPI_ISL_464864\|2020 | ···················· | ···················· | ···················· | ···················· | ···················· | ···················· | |
| D | 8063 | hCoV-19/England/201161657/2020\|EPI_ISL_464869\|2020 | ···················· | ···················· | ···················· | ···················· | ···················· | ···················· | |
| D | 8066 | hCoV-19/England/201180330/2020\|EPI_ISL_464898\|2020 | ···················· | ···················· | ···················· | ···················· | ···················· | ·N···········N······ | |
| D | 8078 | hCoV-19/Spain/CN-IBV-002603/2020\|EPI_ISL_467086\|20 | ···················· | ···················· | ···················· | ···················· | ···················· | ···················· | |
| D | 8081 | hCoV-19/Spain/CN-IBV-002605/2020\|EPI_ISL_467089\|20 | ···················· | ···················· | ···················· | ···················· | ···················· | ···················· | |
| D | 8082 | hCoV-19/Wales/PHWC-1645DD/2020\|EPI_ISL_474201\|2020 | ···················· | ···················· | ···················· | ···················· | ···················· | ···················· | |
| D | 8083 | hCoV-19/Wales/PHWC-1645EC/2020\|EPI_ISL_474202\|2020 | ···················· | ···················· | ···················· | ···················· | ···················· | ···················· | |
| D | 8084 | hCoV-19/Wales/PHWC-164607/2020\|EPI_ISL_474204\|2020 | ···················· | ···················· | ···················· | ···················· | ···················· | ···················· | |
| D | 8085 | hCoV-19/Wales/PHWC-164616/2020\|EPI_ISL_474205\|2020 | ···················· | ···················· | ···················· | ···················· | ···················· | ···················· | |
| D | 8086 | hCoV-19/Wales/PHWC-164625/2020\|EPI_ISL_474206\|2020 | ···················· | ···················· | ···················· | ···················· | ···················· | ···················· | |

| | | | | | | | | |
|---|---|---|---|---|---|---|---|---|
| D | 8087 hCoV-19/Wales/PHWC-164643/2020\|EPI_ISL_474208\|2020 | .................. | .................. | .................. | .................. | .................. | .................. | |
| D | 8088 hCoV-19/Wales/PHWC-164652/2020\|EPI_ISL_474209\|2020 | .................. | .................. | .................. | .................. | .................. | .................. | |
| D | 8089 hCoV-19/Wales/PHWC-164661/2020\|EPI_ISL_474210\|2020 | .................. | .................. | .................. | .................. | .................. | .................. | |
| D | 8090 hCoV-19/Wales/PHWC-164670/2020\|EPI_ISL_474211\|2020 | .................. | .................. | .................. | .................. | .................. | .................. | |
| D | 8091 hCoV-19/Wales/PHWC-29B00/2020\|EPI_ISL_474240\|2020- | .................. | .................. | .................. | .................. | .................. | .................. | |
| D | 8092 hCoV-19/Wales/PHWC-29D19/2020\|EPI_ISL_474248\|2020- | .................. | .................. | .................. | .................. | .................. | .................. | |
| D | 8093 hCoV-19/Wales/PHWC-29E9E/2020\|EPI_ISL_474250\|2020- | .................. | .................. | .................. | .................. | .................. | .................. | |
| D | 8094 hCoV-19/Wales/PHWC-29EE9/2020\|EPI_ISL_474253\|2020- | .................. | .................. | .................. | .................. | .................. | .................. | |
| D | 8095 hCoV-19/Wales/PHWC-29EF8/2020\|EPI_ISL_474254\|2020- | .................. | .................. | .................. | .................. | .................. | .................. | |
| D | 8096 hCoV-19/England/SHEF-D277B/2020\|EPI_ISL_475504\|202 | .................. | .................. | .................. | .................. | .................. | .................. | |
| D | 8097 hCoV-19/England/SHEF-D30AC/2020\|EPI_ISL_475507\|202 | .................. | .................. | .................. | .................. | .................. | .................. | |
| D | 8098 hCoV-19/England/SHEF-D3297/2020\|EPI_ISL_475508\|202 | .................. | .................. | .................. | .................. | .................. | .................. | |
| D | 8099 hCoV-19/England/SHEF-D333A/2020\|EPI_ISL_475509\|202 | .................. | .................. | .................. | .................. | .................. | .................. | |
| D | 8100 hCoV-19/Sweden/20-14131/2020\|EPI_ISL_475511\|2020-0 | .................. | .................. | .................. | .................. | .................. | .................. | |
| D | 8101 hCoV-19/Sweden/20-14137/2020\|EPI_ISL_475512\|2020-0 | .................. | .................. | .................. | .................. | .................. | .................. | |
| D | 8102 hCoV-19/Sweden/20-14178/2020\|EPI_ISL_475513\|2020-0 | .................. | .................. | .................. | .................. | .................. | .................. | |
| D | 8103 hCoV-19/Sweden/20-14387/2020\|EPI_ISL_475514\|2020-0 | .................. | .................. | .................. | .................. | .................. | .................. | |
| D | 8104 hCoV-19/Sweden/20-14509/2020\|EPI_ISL_475515\|2020-0 | .................. | .................. | .................. | .................. | .................. | .................. | |
| D | 8105 hCoV-19/Sweden/20-14518/2020\|EPI_ISL_475516\|2020-0 | .................. | .................. | .................. | .................. | .................. | .................. | |
| D | 8106 hCoV-19/Sweden/20-14519/2020\|EPI_ISL_475517\|2020-0 | .................. | .................. | .................. | .................. | .................. | .................. | |
| D | 8107 hCoV-19/Sweden/20-14679/2020\|EPI_ISL_475518\|2020-0 | .................. | .................. | .................. | .................. | .................. | .................. | |
| D | 8108 hCoV-19/Sweden/20-14682/2020\|EPI_ISL_475519\|2020-0 | .................. | .................. | .................. | .................. | .................. | .................. | |
| D | 8109 hCoV-19/Sweden/20-14734/2020\|EPI_ISL_475520\|2020-0 | .................. | .................. | .................. | .................. | .................. | .................. | |
| D | 8110 hCoV-19/Sweden/20-14828/2020\|EPI_ISL_475521\|2020-0 | .................. | .................. | .................. | .................. | .................. | .................. | |
| D | 8111 hCoV-19/Sweden/20-14937/2020\|EPI_ISL_475522\|2020-0 | .................. | .................. | .................. | .................. | .................. | .................. | |
| D | 8112 hCoV-19/Sweden/20-14941/2020\|EPI_ISL_475523\|2020-0 | .................. | .................. | .................. | .................. | .................. | .................. | |
| D | 8113 hCoV-19/Sweden/20-14985/2020\|EPI_ISL_475524\|2020-0 | .................. | .................. | .................. | .................. | .................. | .................. | |
| D | 8114 hCoV-19/Sweden/20-19194/2020\|EPI_ISL_475525\|2020-0 | .................. | .................. | .................. | .................. | .................. | .................. | |
| D | 8115 hCoV-19/Sweden/20-51516/2020\|EPI_ISL_475526\|2020-0 | .................. | .................. | .................. | .................. | .................. | .................. | |
| D | 8116 hCoV-19/Sweden/20-51753/2020\|EPI_ISL_475527\|2020-0 | .................. | .................. | .................. | .................. | .................. | .................. | |
| D | 8117 hCoV-19/Sweden/20-51755/2020\|EPI_ISL_475528\|2020-0 | .................. | .................. | .................. | .................. | .................. | .................. | |
| D | 8118 hCoV-19/Sweden/20-51780/2020\|EPI_ISL_475529\|2020-0 | .................. | .................. | .................. | .................. | .................. | .................. | |
| D | 8119 hCoV-19/Sweden/20-51829/2020\|EPI_ISL_475530\|2020-0 | .................. | .................. | .................. | .................. | .................. | .................. | |
| D | 8120 hCoV-19/Sweden/20-51830/2020\|EPI_ISL_475531\|2020-0 | .................. | .................. | .................. | .................. | .................. | .................. | |
| D | 8121 hCoV-19/Sweden/20-51831/2020\|EPI_ISL_475532\|2020-0 | .................. | .................. | .................. | .................. | .................. | .................. | |
| D | 8122 hCoV-19/Sweden/20-51900/2020\|EPI_ISL_475533\|2020-0 | .................. | .................. | .................. | .................. | .................. | .................. | |
| D | 8123 hCoV-19/Sweden/20-51902/2020\|EPI_ISL_475534\|2020-0 | .................. | .................. | .................. | .................. | .................. | .................. | |
| D | 8124 hCoV-19/Sweden/20-51955/2020\|EPI_ISL_475535\|2020-0 | .................. | .................. | .................. | .................. | .................. | .................. | |
| D | 8125 hCoV-19/Sweden/20-06070/2020\|EPI_ISL_475536\|2020-0 | .................. | .................. | .................. | .................. | .................. | .................. | |
| D | 8126 hCoV-19/Sweden/20-06918/2020\|EPI_ISL_475537\|2020-0 | .................. | .................. | .................. | .................. | .................. | .................. | |
| D | 8127 hCoV-19/Sweden/20-07012/2020\|EPI_ISL_475538\|2020-0 | .................. | .................. | .................. | .................. | .................. | .................. | |
| D | 8128 hCoV-19/Sweden/20-07017/2020\|EPI_ISL_475539\|2020-0 | .................. | .................. | .................. | .................. | .................. | .................. | |
| D | 8129 hCoV-19/Sweden/20-07382/2020\|EPI_ISL_475540\|2020-0 | .................. | .................. | .................. | .................. | .................. | .................. | |
| D | 8130 hCoV-19/Sweden/20-14071/2020\|EPI_ISL_475541\|2020-0 | .................. | .................. | .................. | .................. | .................. | .................. | |
| D | 8131 hCoV-19/Sweden/20-51337/2020\|EPI_ISL_475542\|2020-0 | .................. | .................. | .................. | .................. | .................. | .................. | |
| D | 8132 hCoV-19/Sweden/20-14611/2020\|EPI_ISL_475566\|2020-0 | .................. | .................. | .................. | .................. | .................. | .................. | |
| D | 8133 hCoV-19/Bangladesh/JUST-GC15.09/2020\|EPI_ISL_47557 | .................. | .................. | .................. | .................. | .................. | .................. | |
| D | 8134 hCoV-19/Bangladesh/JUST-GC40.38/2020\|EPI_ISL_47557 | .................. | .................. | .................. | .................. | .................. | .................. | |
| D | 8135 hCoV-19/England/IC19/2020\|EPI_ISL_475572\|2020-03-1 | .................. | .................. | .................. | .................. | .................. | .................. | |
| D | 8136 hCoV-19/Bangladesh/JUST-GC40.86/2020\|EPI_ISL_47557 | .................. | .................. | .................. | .................. | .................. | .................. | |
| D | 8137 hCoV-19/USA/CA-CSMC10/2020\|EPI_ISL_475574\|2020-03- | .................. | .................. | .................. | .................. | .................. | .................. | |
| D | 8138 hCoV-19/USA/CA-CSMC101/2020\|EPI_ISL_475576\|2020-04 | .................. | .................. | .................. | .................. | .................. | .................. | |
| D | 8139 hCoV-19/USA/CA-CSMC102/2020\|EPI_ISL_475577\|2020-04 | .................. | .................. | .................. | .................. | .................. | .................. | |
| D | 8140 hCoV-19/USA/CA-CSMC103/2020\|EPI_ISL_475578\|2020-03 | .................. | .................. | .................. | .................. | .................. | .................. | |
| D | 8141 hCoV-19/USA/CA-CSMC104/2020\|EPI_ISL_475579\|2020-04 | .................. | .................. | .................. | .................. | .................. | .................. | |
| D | 8142 hCoV-19/USA/CA-CSMC105/2020\|EPI_ISL_475580\|2020-04 | .................. | .................. | .................. | .................. | .................. | .................. | |
| D | 8143 hCoV-19/USA/CA-CSMC107/2020\|EPI_ISL_475582\|2020-04 | .................. | .................. | .................. | .................. | .................. | .................. | |
| D | 8144 hCoV-19/USA/CA-CSMC108/2020\|EPI_ISL_475583\|2020-04 | .................. | .................. | .................. | .................. | .................. | .................. | |
| D | 8145 hCoV-19/USA/CA-CSMC11/2020\|EPI_ISL_475585\|2020-04- | .................. | .................. | .................. | .................. | .................. | .................. | |
| D | 8146 hCoV-19/Hong Kong/VM20003443/2020\|EPI_ISL_476801\|2 | .................. | .................. | .................. | .................. | .................. | .................. | |
| D | 8147 hCoV-19/Hong Kong/VM20003442/2020\|EPI_ISL_476802\|2 | .................. | .................. | .................. | .................. | .................. | .................. | |
| D | 8148 hCoV-19/Hong Kong/VB20050000/2020\|EPI_ISL_476803\|2 | .................. | .................. | .................. | .................. | .................. | .................. | |
| D | 8149 hCoV-19/Hong Kong/VM20004178/2020\|EPI_ISL_476804\|2 | .................. | .................. | .................. | .................. | .................. | .................. | |
| D | 8150 hCoV-19/Benin/1950/2020\|EPI_ISL_476830\|2020-04-15 | .................. | .................. | .................. | .................. | .................. | .................. | |
| D | 8151 hCoV-19/Benin/2012/2020\|EPI_ISL_476831\|2020-04-15 | .................. | .................. | .................. | .................. | .................. | .................. | |
| D | 8152 hCoV-19/Turkey/KOU-MG/2020\|EPI_ISL_476832\|2020-04- | .................. | .................. | .................. | .................. | .................. | .................. | |
| D | 8153 hCoV-19/Benin/461/2020\|EPI_ISL_476833\|2020-03-17 | .................. | .................. | .................. | .................. | .................. | .................. | |
| D | 8154 hCoV-19/Benin/314/2020\|EPI_ISL_476834\|2020-03-17 | .................. | .................. | .................. | .................. | .................. | .................. | |
| D | 8155 hCoV-19/India/MP-DRDE 530/2020\|EPI_ISL_476840\|2020 | .................. | .................. | .................. | .................. | ...............C.. | .................. | 1 |
| D | 8156 hCoV-19/India/MP-DRDE 4764/2020\|EPI_ISL_476842\|202 | .................. | .................. | .................. | .................. | .................. | .................. | |
| D | 8157 hCoV-19/India/MP-DRDE4001/2020\|EPI_ISL_476844\|2020 | .................. | .................. | .................. | .................. | .................. | .................. | |
| D | 8158 hCoV-19/India/MP-DRDE 197/2020\|EPI_ISL_476846\|2020 | .................. | .................. | .................. | .................. | .................. | .................. | |
| D | 8159 hCoV-19/India/MP-DRDE 759/2020\|EPI_ISL_476848\|2020 | .................. | .................. | .................. | .................. | .................. | .................. | |
| D | 8160 hCoV-19/India/MP-DRDE 1794/2020\|EPI_ISL_476849\|202 | .................. | .................. | .................. | .................. | .................. | .................. | |
| D | 8161 hCoV-19/India/MP-DRDE 2429/2020\|EPI_ISL_476850\|202 | .................. | .................. | .................. | .................. | .................. | .................. | |
| D | 8162 hCoV-19/India/MP-DRDE 2497/2020\|EPI_ISL_476852\|202 | .................. | .................. | .................. | .................. | .................. | .................. | |
| D | 8163 hCoV-19/India/MP-DRDE-2498/2020\|EPI_ISL_476853\|202 | .................. | .................. | .................. | .................. | .................. | .................. | |
| D | 8164 hCoV-19/India/MP-DRDE-3302/2020\|EPI_ISL_476854\|202 | .................. | .................. | .................. | .................. | .................. | .................. | |

| | | | | | | | | |
|---|---|---|---|---|---|---|---|---|
| D | 8165 | hCoV-19/India/GJ-GBRC222a/2020\|EPI_ISL_476855\|2020 | ···················· | ·A·················· | ···················· | ···················· | ···················· | ···A················ |
| D | 8166 | hCoV-19/India/GJ-GBRC222b/2020\|EPI_ISL_476856\|2020 | ···················· | ·A·················· | ···················· | ···················· | ···················· | ···A················ |
| D | 8167 | hCoV-19/India/GJ-GBRC223a/2020\|EPI_ISL_476857\|2020 | ···················· | ·A·················· | ···················· | ···················· | ···················· | ···A················ |
| D | 8168 | hCoV-19/India/GJ-GBRC223b/2020\|EPI_ISL_476858\|2020 | ···················· | ·A·················· | ···················· | ···················· | ···················· | ···A················ |
| D | 8169 | hCoV-19/India/GJ-GBRC224a/2020\|EPI_ISL_476859\|2020 | ···················· | ·A·················· | ···················· | ···················· | ···················· | ···A················ |
| D | 8170 | hCoV-19/India/GJ-GBRC224b/2020\|EPI_ISL_476860\|2020 | ···················· | ·A·················· | ···················· | ···················· | ···················· | ···A················ |
| D | 8171 | hCoV-19/India/GJ-GBRC225a/2020\|EPI_ISL_476861\|2020 | ···················· | ·A·················· | ···················· | ···················· | ···················· | ···A················ |
| D | 8172 | hCoV-19/India/GJ-GBRC225b/2020\|EPI_ISL_476862\|2020 | ···················· | ·A·················· | ···················· | ···················· | ···················· | ···A················ |
| D | 8173 | hCoV-19/India/GJ-GBRC226/2020\|EPI_ISL_476863\|2020- | ···················· | ·A·················· | ···················· | ···················· | ···················· | ···A················ |
| D | 8174 | hCoV-19/India/GJ-GBRC227/2020\|EPI_ISL_476864\|2020- | ···················· | ·A·················· | ···················· | ···················· | ···················· | ···A················ |
| D | 8175 | hCoV-19/India/GJ-GBRC228a/2020\|EPI_ISL_476865\|2020 | ···················· | ·A·················· | ···················· | ···················· | ···················· | ···A················ |
| D | 8176 | hCoV-19/India/GJ-GBRC228b/2020\|EPI_ISL_476866\|2020 | ···················· | ·A·················· | ···················· | ···················· | ···················· | ···A················ |
| D | 8177 | hCoV-19/India/GJ-GBRC229a/2020\|EPI_ISL_476867\|2020 | ···················· | ·A·················· | ···················· | ···················· | ···················· | ···A················ |
| D | 8178 | hCoV-19/India/GJ-GBRC229b/2020\|EPI_ISL_476868\|2020 | ···················· | ·A·················· | ···················· | ···················· | ···················· | ···A················ |
| D | 8179 | hCoV-19/India/GJ-GBRC230/2020\|EPI_ISL_476869\|2020- | ···················· | ···················· | ···················· | ···················· | ···················· | ···················· |
| D | 8180 | hCoV-19/India/GJ-GBRC231a/2020\|EPI_ISL_476870\|2020 | ···················· | ·A·················· | ···················· | ···················· | ···················· | ···A················ |
| D | 8181 | hCoV-19/India/GJ-GBRC231b/2020\|EPI_ISL_476871\|2020 | ···················· | ·A·················· | ···················· | ···················· | ···················· | ···A················ |
| D | 8182 | hCoV-19/India/GJ-GBRC232a/2020\|EPI_ISL_476872\|2020 | ···················· | ·A·················· | ···················· | ···················· | ···················· | ···A················ |
| D | 8183 | hCoV-19/India/GJ-GBRC232b/2020\|EPI_ISL_476873\|2020 | ···················· | ·A·················· | ···················· | ···················· | ···················· | ···A················ |
| D | 8184 | hCoV-19/USA/SC-PRG493/2020\|EPI_ISL_476897\|2020-05- | ···················· | ·········A·········· | ···················· | ···················· | ···················· | ···················· |
| D | 8185 | hCoV-19/England/OXON-B1AD3/2020\|EPI_ISL_479070\|202 | ···················· | ···················· | ···················· | ···················· | ···················· | ···················· |
| D | 8186 | hCoV-19/England/OXON-B1AE2/2020\|EPI_ISL_479071\|202 | ···················· | ···················· | ···················· | ···················· | ···················· | ···················· |
| D | 8187 | hCoV-19/England/OXON-B1B1C/2020\|EPI_ISL_479072\|202 | ···················· | ···················· | ···················· | ···················· | ···················· | ···················· |
| D | 8188 | hCoV-19/England/OXON-B1B3A/2020\|EPI_ISL_479073\|202 | ···················· | ···················· | ···················· | ···················· | ···················· | ···················· |
| D | 8189 | hCoV-19/England/OXON-B1B49/2020\|EPI_ISL_479074\|202 | ···················· | ···················· | ···················· | ···················· | ···················· | ···················· |
| D | 8190 | hCoV-19/England/OXON-B1B67/2020\|EPI_ISL_479076\|202 | ···················· | ···················· | ···················· | ···················· | ···················· | ···················· |
| D | 8191 | hCoV-19/England/OXON-B1B76/2020\|EPI_ISL_479077\|202 | ···················· | ···················· | ···················· | ···················· | ···················· | ···················· |
| D | 8192 | hCoV-19/England/OXON-B1BFE/2020\|EPI_ISL_479079\|202 | ···················· | ···················· | ···················· | ···················· | ···················· | ···················· |
| D | 8193 | hCoV-19/USA/WA-S1010/2020\|EPI_ISL_463500\|2020-04-2 | ···················· | ···················· | ···················· | ···················· | ···················· | ···················· |
| D | 8194 | hCoV-19/USA/WA-S1011/2020\|EPI_ISL_463501\|2020-04-2 | ···················· | ···················· | ···················· | ···················· | ···················· | ···················· |
| D | 8195 | hCoV-19/USA/WA-S1012/2020\|EPI_ISL_463502\|2020-04-2 | ···················· | ···················· | ···················· | ···················· | ···················· | ···················· |
| D | 8196 | hCoV-19/USA/WA-S1013/2020\|EPI_ISL_463503\|2020-04-3 | ···················· | ···················· | ···················· | ···················· | ···················· | ···················· |
| D | 8197 | hCoV-19/USA/WA-S1014/2020\|EPI_ISL_463504\|2020-04-3 | ···················· | ···················· | ···················· | ···················· | ···················· | ···················· |
| D | 8198 | hCoV-19/USA/WA-S1015/2020\|EPI_ISL_463505\|2020-04-3 | ···················· | ···················· | ···················· | ···················· | ···················· | ···················· |
| D | 8199 | hCoV-19/USA/WA-S1016/2020\|EPI_ISL_463506\|2020-04-2 | ···················· | ···················· | ···················· | ···················· | ···················· | ···················· |
| D | 8200 | hCoV-19/USA/WA-S1017/2020\|EPI_ISL_463507\|2020-04-3 | ···················· | ···················· | ···················· | ···················· | ···················· | ···················· |
| D | 8201 | hCoV-19/USA/WA-S1018/2020\|EPI_ISL_463508\|2020-04-3 | ···················· | ···················· | ···················· | ···················· | ···················· | ···················· |
| D | 8202 | hCoV-19/USA/WA-S1019/2020\|EPI_ISL_463509\|2020-04-3 | ···················· | ···················· | ···················· | ···················· | ···················· | ···················· |
| D | 8203 | hCoV-19/USA/WA-S1020/2020\|EPI_ISL_463510\|2020-04-2 | ···················· | ···················· | ···················· | ···················· | ···················· | ···················· |
| D | 8204 | hCoV-19/USA/WA-S1021/2020\|EPI_ISL_463511\|2020-04-3 | ···················· | ···················· | ···················· | ···················· | ···················· | ···················· |
| D | 8205 | hCoV-19/USA/WA-S1022/2020\|EPI_ISL_463512\|2020-04-2 | ···················· | ···················· | ···················· | ···················· | ···················· | ···················· |
| D | 8206 | hCoV-19/USA/WA-S1023/2020\|EPI_ISL_463513\|2020-05-0 | ···················· | ···················· | ···················· | ···················· | ···················· | ···················· |
| D | 8207 | hCoV-19/USA/WA-S1024/2020\|EPI_ISL_463514\|2020-05-0 | ···················· | ···················· | ···················· | ···················· | ···················· | ···················· |
| D | 8208 | hCoV-19/USA/WA-S1025/2020\|EPI_ISL_463515\|2020-05-0 | ···················· | ···················· | ···················· | ···················· | ···················· | ···················· |
| D | 8209 | hCoV-19/USA/WA-S1026/2020\|EPI_ISL_463516\|2020-04-3 | ···················· | ···················· | ···················· | ···················· | ···················· | ···················· |
| D | 8210 | hCoV-19/USA/WA-S1027/2020\|EPI_ISL_463517\|2020-05-0 | ···················· | ···················· | ···················· | ···················· | ···················· | ···················· |
| D | 8211 | hCoV-19/USA/WA-S1028/2020\|EPI_ISL_463518\|2020-05-0 | ···················· | ···················· | ···················· | ···················· | ···················· | ···················· |
| D | 8212 | hCoV-19/USA/WA-S1029/2020\|EPI_ISL_463519\|2020-05-0 | ···················· | ···················· | ···················· | ···················· | ···················· | ···················· |
| D | 8217 | hCoV-19/England/201161180/2020\|EPI_ISL_464817\|2020 | ···················· | ···················· | ···················· | ···················· | ···················· | ···················· |
| D | 8221 | hCoV-19/England/201161204/2020\|EPI_ISL_464822\|2020 | ···················· | ···················· | ···················· | ···················· | ···················· | ···················· |
| D | 8222 | hCoV-19/England/201161219/2020\|EPI_ISL_464824\|2020 | ···················· | ···················· | ···················· | ···················· | ···················· | ···················· |
| D | 8224 | hCoV-19/England/201161300/2020\|EPI_ISL_464830\|2020 | ···················· | ···················· | ···················· | ···················· | ···················· | ···················· |
| D | 8227 | hCoV-19/England/201161375/2020\|EPI_ISL_464835\|2020 | ···················· | ···················· | ···················· | ···················· | ···················· | ···················· |
| D | 8229 | hCoV-19/England/201161424/2020\|EPI_ISL_464838\|2020 | ···················· | ···················· | ···················· | ···················· | ···················· | ···················· |
| D | 8231 | hCoV-19/England/201161452/2020\|EPI_ISL_464841\|2020 | ···················· | ···················· | ···················· | ···················· | ···················· | N··················· |
| D | 8233 | hCoV-19/England/201161462/2020\|EPI_ISL_464843\|2020 | ···················· | ···················· | ···················· | ···················· | ···················· | ···················· |
| D | 8234 | hCoV-19/England/201161466/2020\|EPI_ISL_464844\|2020 | ···················· | ···················· | ···················· | ···················· | ···················· | ···················· |
| D | 8235 | hCoV-19/England/201161484/2020\|EPI_ISL_464849\|2020 | ···················· | ···················· | ···················· | ···················· | ···················· | ···················· |
| D | 8237 | hCoV-19/Switzerland/ZH-ETHZ-130006/2020\|EPI_ISL_46 | ···················· | ···················· | ···················· | ···················· | ···················· | ···················· |
| D | 8239 | hCoV-19/Switzerland/VD-ETHZ-130008/2020\|EPI_ISL_46 | ···················· | ·A·················· | ···················· | ···················· | ···················· | ···················· |
| D | 8241 | hCoV-19/India/GJ-GBRC181b/2020\|EPI_ISL_467050\|2020 | ···················· | ·A·················· | ···················· | ···················· | ···················· | ···A················ |
| D | 8242 | hCoV-19/India/GJ-GBRC182a/2020\|EPI_ISL_467051\|2020 | ···················· | ·A·················· | ···················· | ···················· | ···················· | ···A················ |
| D | 8243 | hCoV-19/India/GJ-GBRC182b/2020\|EPI_ISL_467052\|2020 | ···················· | ···················· | ···················· | ···················· | ···················· | ···················· |
| D | 8244 | hCoV-19/India/GJ-GBRC183a/2020\|EPI_ISL_467053\|2020 | ···················· | ·A·················· | ···················· | ···················· | ···················· | ···A················ |
| D | 8245 | hCoV-19/India/GJ-GBRC183b/2020\|EPI_ISL_467054\|2020 | ···················· | ·A·················· | ···················· | ···················· | ···················· | ···A················ |
| D | 8251 | hCoV-19/Spain/AN-IBV-002910/2020\|EPI_ISL_467062\|20 | ···················· | ···················· | ···················· | ···················· | ···················· | ···················· |
| D | 8262 | hCoV-19/Spain/CN-IBV-002595/2020\|EPI_ISL_467090\|20 | ···················· | ···················· | ···················· | ···················· | ···················· | ···················· |
| D | 8264 | hCoV-19/Spain/PV-IBV-001464/2020\|EPI_ISL_467092\|20 | ···················· | ···················· | ···················· | ···················· | ···················· | ···················· |
| D | 8265 | hCoV-19/Spain/PV-IBV-001514/2020\|EPI_ISL_467093\|20 | ···················· | ···················· | ···················· | ···················· | ···················· | ···················· |
| D | 8266 | hCoV-19/Spain/PV-IBV-001551/2020\|EPI_ISL_467094\|20 | ···················· | ···················· | ···················· | ···················· | ···················· | ···················· |
| D | 8267 | hCoV-19/Spain/PV-IBV-001434/2020\|EPI_ISL_467095\|20 | ···················· | ···················· | ···················· | ···················· | ···················· | ···················· |
| D | 8268 | hCoV-19/Spain/PV-IBV-001431/2020\|EPI_ISL_467096\|20 | ···················· | ···················· | ···················· | ···················· | ···················· | ···················· |
| D | 8271 | hCoV-19/Spain/PV-IBV-001533/2020\|EPI_ISL_467099\|20 | ···················· | ···················· | ···················· | ···················· | ···················· | ···················· |
| D | 8272 | hCoV-19/Switzerland/ZH-ETHZ-120244/2020\|EPI_ISL_46 | ···················· | ···················· | ···················· | ···················· | ···················· | ···················· |
| D | 8273 | hCoV-19/Brazil/SP-138/2020\|EPI_ISL_468305\|2020-04- | ···················· | ···················· | ···················· | ···················· | ···················· | ···················· |
| D | 8274 | hCoV-19/Brazil/SP-140/2020\|EPI_ISL_468307\|2020-04- | ···················· | ···················· | ···················· | ···················· | ···················· | ···················· |
| D | 8275 | hCoV-19/Brazil/SP-141/2020\|EPI_ISL_468308\|2020-03- | ···················· | ···················· | ···················· | ···················· | ···················· | ···················· |
| D | 8276 | hCoV-19/Brazil/SP-144/2020\|EPI_ISL_468310\|2020-03- | ···················· | ···················· | ···················· | ···················· | ···················· | ···················· |

| | | | | | | | | |
|---|---|---|---|---|---|---|---|---|
| D | 8277 | hCoV-19/Brazil/SP-146/2020\|EPI_ISL_468311\|2020-03- | ···················· | ···················· | ···················· | ···················· | ···················· | ···················· |
| D | 8278 | hCoV-19/Brazil/SP-147/2020\|EPI_ISL_468312\|2020-03- | ···················· | ···················· | ···················· | ···················· | ···················· | ···················· |
| D | 8279 | hCoV-19/Brazil/SP-148/2020\|EPI_ISL_468313\|2020-03- | ···················· | ···················· | ···················· | ···················· | ···················· | ···················· |
| D | 8280 | hCoV-19/Brazil/SP-149/2020\|EPI_ISL_468314\|2020-03- | ···················· | ···················· | ···················· | ···················· | ···················· | ···················· |
| D | 8281 | hCoV-19/Brazil/SP-500/2020\|EPI_ISL_468315\|2020-04- | ···················· | ···················· | ···················· | ···················· | ···················· | ···················· |
| D | 8282 | hCoV-19/Brazil/SP-504/2020\|EPI_ISL_468316\|2020-04- | ···················· | ···················· | ···················· | ···················· | ···················· | ···················· |
| D | 8283 | hCoV-19/Brazil/SP-505/2020\|EPI_ISL_468318\|2020-04- | ···················· | ···················· | ···················· | ···················· | ···················· | ···················· |
| D | 8284 | hCoV-19/Brazil/SP-506/2020\|EPI_ISL_468319\|2020-04- | ···················· | ···················· | ···················· | ···················· | ···················· | ···················· |
| D | 8285 | hCoV-19/Brazil/SP-508/2020\|EPI_ISL_468320\|2020-04- | ···················· | ···················· | ···················· | ···················· | ···················· | ···················· |
| D | 8286 | hCoV-19/Brazil/SP-516/2020\|EPI_ISL_468321\|2020-04- | ···················· | ···················· | ···················· | ···················· | ···················· | ···················· |
| D | 8287 | hCoV-19/USA/CA-CZB-1400/2020\|EPI_ISL_468380\|2020-0 | ···R················ | ···················· | ···················· | ···················· | ···················· | ···················· |
| D | 8288 | hCoV-19/USA/CA-CZB-1402/2020\|EPI_ISL_468382\|2020-0 | ···················· | ···················· | ···················· | ···················· | ···················· | ···················· |
| D | 8289 | hCoV-19/USA/CA-CZB-1404/2020\|EPI_ISL_468383\|2020-0 | ···················· | ···················· | ···················· | ···················· | ···················· | ···················· |
| D | 8290 | hCoV-19/USA/CA-CZB-1410/2020\|EPI_ISL_468385\|2020-0 | ···················· | ···················· | ···················· | ···················· | ···················· | ···················· |
| D | 8291 | hCoV-19/USA/CA-CZB-1412/2020\|EPI_ISL_468387\|2020-0 | ···················· | ···················· | ···················· | ···················· | ···················· | ···················· |
| D | 8292 | hCoV-19/USA/CA-CZB-1413/2020\|EPI_ISL_468388\|2020-0 | ···················· | ···················· | ···················· | ···················· | ···················· | ···················· |
| D | 8293 | hCoV-19/USA/CA-CZB-1414/2020\|EPI_ISL_468389\|2020-0 | ···················· | ···················· | ···················· | ···················· | ···················· | ···················· |
| D | 8294 | hCoV-19/USA/CA-CZB-1415/2020\|EPI_ISL_468390\|2020-0 | ···················· | ···················· | ···················· | ···················· | ···················· | ···················· |
| D | 8295 | hCoV-19/USA/CA-CZB-1417/2020\|EPI_ISL_468391\|2020-0 | ···················· | ···················· | ···················· | ···················· | ···················· | ···················· |
| D | 8296 | hCoV-19/USA/CA-CZB-1418/2020\|EPI_ISL_468392\|2020-0 | ···················· | ···················· | ···················· | ···················· | ···················· | ···················· |
| D | 8297 | hCoV-19/USA/CA-CZB-1420/2020\|EPI_ISL_468393\|2020-0 | ···················· | ···················· | ···················· | ···················· | ···················· | ···················· |
| D | 8298 | hCoV-19/USA/CA-CZB-1421/2020\|EPI_ISL_468394\|2020-0 | ···················· | ···················· | ···················· | ···················· | ···················· | ···················· |
| D | 8299 | hCoV-19/USA/CA-CZB-1423/2020\|EPI_ISL_468395\|2020-0 | ···················· | ···················· | ···················· | ···················· | ···················· | ···················· |
| D | 8300 | hCoV-19/USA/CA-CZB-1424/2020\|EPI_ISL_468396\|2020-0 | ···················· | ···················· | ···················· | ···················· | ···················· | ···················· |
| D | 8301 | hCoV-19/USA/CA-CZB-1425/2020\|EPI_ISL_468397\|2020-0 | ···················· | ···················· | ···················· | ···················· | ···················· | ···················· |
| D | 8302 | hCoV-19/USA/CA-CZB-1427/2020\|EPI_ISL_468398\|2020-0 | ···················· | ···················· | ···················· | ···················· | ···················· | ···················· |
| D | 8303 | hCoV-19/USA/CA-CZB-1428/2020\|EPI_ISL_468399\|2020-0 | ···················· | ···················· | ···················· | ···················· | ···················· | ···················· |
| D | 8304 | hCoV-19/England/BRIS-127D38/2020\|EPI_ISL_469630\|20 | ···················· | ···················· | ···················· | ···················· | ···················· | ···················· |
| D | 8305 | hCoV-19/England/BRIS-12713C/2020\|EPI_ISL_469632\|20 | ···················· | ···················· | ···················· | ···················· | ···················· | ···················· |
| D | 8306 | hCoV-19/England/BRIS-127855/2020\|EPI_ISL_469634\|20 | ···················· | ···················· | ···················· | ···················· | ···················· | ···················· |
| D | 8307 | hCoV-19/England/BRIS-1276B5/2020\|EPI_ISL_469636\|20 | ···················· | ···················· | ···················· | ···················· | ···················· | ···················· |
| D | 8308 | hCoV-19/England/BRIS-127248/2020\|EPI_ISL_469637\|20 | ···················· | ···················· | ···················· | ···················· | ···················· | ···················· |
| D | 8309 | hCoV-19/England/BRIS-128353/2020\|EPI_ISL_469638\|20 | ···················· | ···················· | ···················· | ···················· | ···················· | ···················· |
| D | 8310 | hCoV-19/England/BRIS-127C1D/2020\|EPI_ISL_469639\|20 | ···················· | ···················· | ···················· | ···················· | ···················· | ···················· |
| D | 8311 | hCoV-19/England/BRIS-12705D/2020\|EPI_ISL_469641\|20 | ···················· | ···················· | ···················· | ···················· | ···················· | ···················· |
| D | 8312 | hCoV-19/England/BRIS-127512/2020\|EPI_ISL_469643\|20 | ···················· | ···················· | ···················· | ···················· | ···················· | ···················· |
| D | 8313 | hCoV-19/England/BRIS-127CA4/2020\|EPI_ISL_469644\|20 | ···················· | ···················· | ···················· | ···················· | ···················· | ···················· |
| D | 8314 | hCoV-19/England/BRIS-1278FB/2020\|EPI_ISL_469645\|20 | ···················· | ···················· | ···················· | ···················· | ···················· | ···················· |
| D | 8315 | hCoV-19/England/BRIS-127178/2020\|EPI_ISL_469649\|20 | ···················· | ···················· | ···················· | ···················· | ···················· | ···················· |
| D | 8316 | hCoV-19/England/BRIS-126FCA/2020\|EPI_ISL_469651\|20 | ···················· | ···················· | ···················· | ···················· | ···················· | ···················· |
| D | 8317 | hCoV-19/England/BRIS-1283CC/2020\|EPI_ISL_469652\|20 | ···················· | ···················· | ···················· | ···················· | ···················· | ···················· |
| D | 8318 | hCoV-19/England/BRIS-127E62/2020\|EPI_ISL_469654\|20 | ···················· | ···················· | ···················· | ···················· | ···················· | ···················· |
| D | 8319 | hCoV-19/England/BRIS-1271A5/2020\|EPI_ISL_469657\|20 | ···················· | ···················· | ···················· | ···················· | ···················· | ···················· |
| D | 8320 | hCoV-19/England/BRIS-1283DB/2020\|EPI_ISL_469659\|20 | ···················· | ···················· | ···················· | ···················· | ···················· | ···················· |
| D | 8321 | hCoV-19/England/BRIS-127AF5/2020\|EPI_ISL_469661\|20 | ···················· | ···················· | ···················· | ···················· | ···················· | ···················· |
| D | 8322 | hCoV-19/England/BRIS-127433/2020\|EPI_ISL_469663\|20 | ···················· | ···················· | ···················· | ···················· | ···················· | ···················· |
| D | 8323 | hCoV-19/England/BRIS-12849C/2020\|EPI_ISL_469664\|20 | ···················· | ···················· | ···················· | ···················· | ···················· | ···················· |
| D | 8324 | hCoV-19/England/BRIS-1272A2/2020\|EPI_ISL_469665\|20 | ···················· | ···················· | ···················· | ···················· | ···················· | ···················· |
| D | 8325 | hCoV-19/England/BRIS-1281E0/2020\|EPI_ISL_469666\|20 | ···················· | ···················· | ···················· | ···················· | ···················· | ···················· |
| D | 8326 | hCoV-19/England/BRIS-126F24/2020\|EPI_ISL_469667\|20 | ···················· | ····A··············· | ···················· | ···················· | ···················· | ···················· |
| D | 8327 | hCoV-19/England/BRIS-127D65/2020\|EPI_ISL_469668\|20 | ···················· | ···················· | ···················· | ···················· | ···················· | ···················· |
| D | 8328 | hCoV-19/USA/UT-01850/2020\|EPI_ISL_470656\|2020-04-1 | ···················· | ···················· | ···················· | ···················· | ···················· | ···················· |
| D | 8329 | hCoV-19/USA/UT-01852/2020\|EPI_ISL_470657\|2020-04-1 | ···················· | ···················· | ···················· | ···················· | ···················· | ···················· |
| D | 8330 | hCoV-19/USA/UT-01854/2020\|EPI_ISL_470659\|2020-04-1 | ···················· | ···················· | ···················· | ···················· | ···················· | ···················· |
| D | 8331 | hCoV-19/USA/UT-02033/2020\|EPI_ISL_470660\|2020-04-1 | ···················· | ···················· | ···················· | ···················· | ···················· | ···················· |
| D | 8332 | hCoV-19/USA/UT-02036/2020\|EPI_ISL_470662\|2020-04-1 | ···················· | ···················· | ···················· | ···················· | ···················· | ···················· |
| D | 8333 | hCoV-19/USA/UT-02041/2020\|EPI_ISL_470664\|2020-04-1 | ···················· | ···················· | ···················· | ···················· | ···················· | ···················· |
| D | 8334 | hCoV-19/USA/UT-02044/2020\|EPI_ISL_470666\|2020-04-1 | ···················· | ···················· | ···················· | ···················· | ···················· | ···················· |
| D | 8335 | hCoV-19/USA/UT-02047/2020\|EPI_ISL_470668\|2020-04-2 | ···················· | ···················· | ···················· | ···················· | ···················· | ···················· |
| D | 8336 | hCoV-19/USA/UT-02051/2020\|EPI_ISL_470669\|2020-04-2 | ···················· | ···················· | ···················· | ···················· | ···················· | ···················· |
| D | 8337 | hCoV-19/USA/UT-02054/2020\|EPI_ISL_470670\|2020-04-2 | ···················· | ···················· | ···················· | ···················· | ···················· | ···················· |
| D | 8338 | hCoV-19/USA/UT-02423/2020\|EPI_ISL_470673\|2020-04-2 | ···················· | ···················· | ···················· | ···················· | ···················· | ···················· |
| D | 8339 | hCoV-19/USA/UT-02429/2020\|EPI_ISL_470677\|2020-04-2 | ···················· | ···················· | ···················· | ···················· | ···················· | ···················· |
| D | 8340 | hCoV-19/USA/UT-03822/2020\|EPI_ISL_470678\|2020-04-1 | ···················· | ···················· | ···················· | ···················· | ···················· | ···················· |
| D | 8341 | hCoV-19/USA/UT-01837/2020\|EPI_ISL_470680\|2020-04-1 | ···················· | ···················· | ···················· | ···················· | ···················· | ···················· |
| D | 8342 | hCoV-19/USA/UT-01838/2020\|EPI_ISL_470681\|2020-04-1 | ···················· | ···················· | ···················· | ···················· | ···················· | ···················· |
| D | 8343 | hCoV-19/USA/UT-01841/2020\|EPI_ISL_470683\|2020-04-1 | ···················· | ···················· | ···················· | ···················· | ···················· | ···················· |
| D | 8344 | hCoV-19/USA/UT-01842/2020\|EPI_ISL_470684\|2020-04-1 | ···················· | ···················· | ···················· | ···················· | ···················· | ···················· |
| D | 8345 | hCoV-19/USA/UT-01843/2020\|EPI_ISL_470685\|2020-04-1 | ···················· | ···················· | ···················· | ···················· | ···················· | ···················· |
| D | 8346 | hCoV-19/USA/UT-01844/2020\|EPI_ISL_470686\|2020-04-1 | ···················· | ···················· | ···················· | ···················· | ···················· | ···················· |
| D | 8347 | hCoV-19/USA/UT-01845/2020\|EPI_ISL_470687\|2020-04-1 | ···················· | ···················· | ···················· | ···················· | ···················· | ···················· |
| D | 8348 | hCoV-19/USA/UT-01846/2020\|EPI_ISL_470688\|2020-04-1 | ···················· | ···················· | ···················· | ···················· | ···················· | ···················· |
| D | 8349 | hCoV-19/USA/UT-01870/2020\|EPI_ISL_470694\|2020-04-1 | ···················· | ···················· | ···················· | ···················· | ···················· | ···················· |
| D | 8350 | hCoV-19/USA/UT-01872/2020\|EPI_ISL_470696\|2020-04-1 | ···················· | ···················· | ···················· | ···················· | ···················· | ···················· |
| D | 8351 | hCoV-19/USA/UT-01873/2020\|EPI_ISL_470697\|2020-04-1 | ···················· | ···················· | ···················· | ···················· | ···················· | ···················· |
| D | 8352 | hCoV-19/USA/UT-01875/2020\|EPI_ISL_470699\|2020-04-1 | ···················· | ···················· | ···················· | ···················· | ···················· | ···················· |
| D | 8353 | hCoV-19/England/EXET-13761C/2020\|EPI_ISL_471981\|20 | ···················· | ···················· | ···················· | ···················· | ···················· | ···················· |
| D | 8356 | hCoV-19/England/EXET-137852/2020\|EPI_ISL_471989\|20 | ···················· | ···················· | ···················· | ···················· | ···················· | ···················· |

| | | | | | | | | |
|---|---|---|---|---|---|---|---|---|
| D | 8357 | hCoV-19/England/EXET-13789E/2020\|EPI_ISL_471990\|20 | ···················· | ···················· | ····················· | ···················· | ···················· | ···················· |
| D | 8359 | hCoV-19/England/EXET-1378DA/2020\|EPI_ISL_471992\|20 | ···················· | ···················· | ····················· | ···················· | ···················· | ···················· |
| D | 8360 | hCoV-19/England/LIVE-9F640/2020\|EPI_ISL_471993\|202 | ···················· | ···················· | ····················· | ···················· | ···················· | ···················· |
| D | 8361 | hCoV-19/England/LIVE-9F65F/2020\|EPI_ISL_471994\|202 | ···················· | ···················· | ····················· | ···················· | ···················· | ···················· |
| D | 8362 | hCoV-19/England/LIVE-9F66E/2020\|EPI_ISL_471995\|202 | ···················· | ···················· | ····················· | ···················· | ···················· | ···················· |
| D | 8363 | hCoV-19/England/LIVE-9F67D/2020\|EPI_ISL_471996\|202 | ···················· | ···················· | ····················· | ···················· | ···················· | ···················· |
| D | 8364 | hCoV-19/England/LIVE-9F68C/2020\|EPI_ISL_471997\|202 | ···················· | ···················· | ····················· | ···················· | ···················· | ···················· |
| D | 8365 | hCoV-19/England/LIVE-9F69B/2020\|EPI_ISL_471998\|202 | ···················· | ···················· | ····················· | ···················· | ···················· | ···················· |
| D | 8366 | hCoV-19/England/LIVE-9F6AA/2020\|EPI_ISL_471999\|202 | ···················· | ···················· | ····················· | ···················· | ···················· | ···················· |
| D | 8367 | hCoV-19/Singapore/478/2020\|EPI_ISL_476814\|2020-03- | ···················· | ···················· | ····················· | ···················· | ···················· | ···················· |
| D | 8368 | hCoV-19/Singapore/455nan/2020\|EPI_ISL_476818\|2020- | ···················· | ···················· | ····················· | ···················· | ···················· | ···················· |
| D | 8369 | hCoV-19/Singapore/274nan/2020\|EPI_ISL_476819\|2020- | ···················· | ···················· | ····················· | ···················· | ···················· | ···················· |
| D | 8370 | hCoV-19/Benin/197/2020\|EPI_ISL_476822\|2020-03-15 | ···················· | ···················· | ····················· | ···················· | ···················· | ···················· |
| D | 8371 | hCoV-19/Benin/260/2020\|EPI_ISL_476823\|2020-03-17 | ···················· | ···················· | ····················· | ···················· | ···················· | ···················· |
| D | 8372 | hCoV-19/Benin/501/2020\|EPI_ISL_476824\|2020-03-17 | ···················· | ···················· | ····················· | ···················· | ···················· | ···················· |
| D | 8373 | hCoV-19/Benin/1408/2020\|EPI_ISL_476825\|2020-04-04 | ···················· | ···················· | ····················· | ···················· | ···················· | ···················· |
| D | 8374 | hCoV-19/Benin/1022/2020\|EPI_ISL_476826\|2020-03-25 | ···················· | ···················· | ····················· | ···················· | ···················· | ···················· |
| D | 8375 | hCoV-19/Benin/1092/2020\|EPI_ISL_476827\|2020-03-27 | ···················· | ···················· | ····················· | ···················· | ···················· | ···················· |
| D | 8376 | hCoV-19/Benin/1409/2020\|EPI_ISL_476828\|2020-04-06 | ···················· | ···················· | ····················· | ···················· | ···················· | ···················· |
| D | 8377 | hCoV-19/Benin/843/2020\|EPI_ISL_476829\|2020-03-19 | ···················· | ···················· | ····················· | ···················· | ···················· | ···················· |
| D | 8378 | hCoV-19/England/OXON-B12CC/2020\|EPI_ISL_479001\|202 | ···················· | ·A·················· | ····················· | ···················· | ···················· | ···················· |
| D | 8379 | hCoV-19/England/OXON-B12EA/2020\|EPI_ISL_479003\|202 | ···················· | ···················· | ····················· | ···················· | ···················· | ···················· |
| D | 8380 | hCoV-19/England/OXON-B1332/2020\|EPI_ISL_479005\|202 | ···················· | ···················· | ····················· | ···················· | ···················· | ···················· |
| D | 8381 | hCoV-19/England/OXON-B1341/2020\|EPI_ISL_479006\|202 | ···················· | ···················· | ····················· | ···················· | ···················· | ···················· |
| D | 8382 | hCoV-19/England/OXON-B1350/2020\|EPI_ISL_479007\|202 | ···················· | ···················· | ····················· | ···················· | ···················· | ···················· |
| D | 8383 | hCoV-19/England/OXON-B13F6/2020\|EPI_ISL_479011\|202 | ···················· | ···················· | ····················· | ···················· | ···················· | ···················· |
| D | 8384 | hCoV-19/England/OXON-B1402/2020\|EPI_ISL_479012\|202 | ···················· | ···················· | ····················· | ···················· | ···················· | ···················· |
| D | 8385 | hCoV-19/England/OXON-B1411/2020\|EPI_ISL_479013\|202 | ···················· | ···················· | ····················· | ···················· | ···················· | ···················· |
| D | 8386 | hCoV-19/England/OXON-B1420/2020\|EPI_ISL_479014\|202 | ···················· | ···················· | ····················· | ···················· | ···················· | ···················· |
| D | 8387 | hCoV-19/England/OXON-B145D/2020\|EPI_ISL_479015\|202 | ···················· | ···················· | ····················· | ···················· | ···················· | ···················· |
| D | 8388 | hCoV-19/England/OXON-B147B/2020\|EPI_ISL_479016\|202 | ···················· | ···················· | ····················· | ···················· | ···················· | ···················· |
| D | 8389 | hCoV-19/England/OXON-B148A/2020\|EPI_ISL_479017\|202 | ···················· | ···················· | ····················· | ···················· | ···················· | ···················· |
| D | 8390 | hCoV-19/England/OXON-B1499/2020\|EPI_ISL_479018\|202 | ···················· | ···················· | ····················· | ···················· | ···················· | ···················· |
| D | 8391 | hCoV-19/England/OXON-B14B7/2020\|EPI_ISL_479019\|202 | ···················· | ···················· | ····················· | ···················· | ···················· | ···················· |
| D | 8392 | hCoV-19/England/OXON-B14E4/2020\|EPI_ISL_479022\|202 | ···················· | ···················· | ····················· | ···················· | ···················· | ···················· |
| D | 8393 | hCoV-19/England/OXON-B14F3/2020\|EPI_ISL_479023\|202 | ···················· | ···················· | ····················· | ···················· | ···················· | ···················· |
| D | 8394 | hCoV-19/England/OXON-B151E/2020\|EPI_ISL_479024\|202 | ···················· | ···················· | ····················· | ···················· | ···················· | ···················· |
| D | 8395 | hCoV-19/England/OXON-B160C/2020\|EPI_ISL_479030\|202 | ···················· | ···················· | ····················· | ···················· | ···················· | ···················· |
| D | 8396 | hCoV-19/England/OXON-B162A/2020\|EPI_ISL_479031\|202 | ···················· | ···················· | ····················· | ···················· | ···················· | ···················· |
| D | 8397 | hCoV-19/England/OXON-B1657/2020\|EPI_ISL_479032\|202 | ···················· | ···················· | ····················· | ···················· | ···················· | ···················· |
| D | 8398 | hCoV-19/England/OXON-B1693/2020\|EPI_ISL_479033\|202 | ···················· | ···················· | ····················· | ···················· | ···················· | ···················· |
| D | 8399 | hCoV-19/England/OXON-B16B1/2020\|EPI_ISL_479035\|202 | ···················· | ···················· | ····················· | ···················· | ···················· | ···················· |
| D | 8400 | hCoV-19/England/OXON-B16C0/2020\|EPI_ISL_479036\|202 | ···················· | ·A·················· | ····················· | ···················· | ···················· | ···················· |
| D | 8401 | hCoV-19/England/OXON-B16DF/2020\|EPI_ISL_479037\|202 | ···················· | ···················· | ····················· | ···················· | ···················· | ···················· |
| D | 8402 | hCoV-19/England/OXON-B1709/2020\|EPI_ISL_479038\|202 | ···················· | ···················· | ····················· | ···················· | ···················· | ···················· |
| D | 8403 | hCoV-19/England/OXON-B1736/2020\|EPI_ISL_479040\|202 | ···················· | ···················· | ····················· | ···················· | ···················· | ···················· |
| D | 8404 | hCoV-19/England/OXON-B1754/2020\|EPI_ISL_479041\|202 | ···················· | ···················· | ····················· | ···················· | ···················· | ···················· |
| D | 8405 | hCoV-19/England/OXON-B1763/2020\|EPI_ISL_479042\|202 | ···················· | ···················· | ····················· | ···················· | ···················· | ···················· |
| D | 8406 | hCoV-19/England/OXON-B1772/2020\|EPI_ISL_479043\|202 | ···················· | ···················· | ····················· | ···················· | ···················· | ···················· |
| D | 8407 | hCoV-19/England/OXON-B1781/2020\|EPI_ISL_479044\|202 | ···················· | ·A·················· | ····················· | ···················· | ···················· | ···················· |
| D | 8408 | hCoV-19/England/OXON-B1790/2020\|EPI_ISL_479045\|202 | ···················· | ···················· | ····················· | ···················· | ···················· | ···················· |
| D | 8409 | hCoV-19/England/OXON-B17EB/2020\|EPI_ISL_479046\|202 | ···················· | ···················· | ····················· | ···················· | ···················· | ···················· |
| D | 8410 | hCoV-19/England/OXON-B1806/2020\|EPI_ISL_479047\|202 | ···················· | ···················· | ····················· | ···················· | ···················· | ···················· |
| D | 8411 | hCoV-19/England/OXON-B1824/2020\|EPI_ISL_479048\|202 | ···················· | ···················· | ····················· | ···················· | ···················· | ···················· |
| D | 8412 | hCoV-19/England/OXON-B18CA/2020\|EPI_ISL_479050\|202 | ···················· | ···················· | ····················· | ···················· | ···················· | ···················· |
| D | 8413 | hCoV-19/England/OXON-B18D9/2020\|EPI_ISL_479051\|202 | ···················· | ···················· | ····················· | ···················· | ···················· | ···················· |
| D | 8414 | hCoV-19/England/OXON-B18F7/2020\|EPI_ISL_479052\|202 | ···················· | ···················· | ····················· | ···················· | ···················· | ···················· |
| D | 8415 | hCoV-19/England/OXON-B194F/2020\|EPI_ISL_479054\|202 | ···················· | ···················· | ····················· | ···················· | ···················· | ···················· |
| D | 8416 | hCoV-19/England/OXON-B195E/2020\|EPI_ISL_479055\|202 | ···················· | ···················· | ····················· | ···················· | ···················· | ···················· |
| D | 8417 | hCoV-19/England/OXON-B196D/2020\|EPI_ISL_479056\|202 | ···················· | ···················· | ····················· | ···················· | ···················· | ···················· |
| D | 8418 | hCoV-19/England/OXON-B198B/2020\|EPI_ISL_479057\|202 | ···················· | ·A·················· | ····················· | ···················· | ···················· | ···················· |
| D | 8419 | hCoV-19/England/OXON-B199A/2020\|EPI_ISL_479058\|202 | ···················· | ···················· | ····················· | ···················· | ···················· | ···················· |
| D | 8420 | hCoV-19/England/OXON-B19A9/2020\|EPI_ISL_479059\|202 | ···················· | ···················· | ····················· | ···················· | ···················· | ···················· |
| D | 8421 | hCoV-19/England/OXON-B19C7/2020\|EPI_ISL_479060\|202 | ···················· | ···················· | ····················· | ···················· | ···················· | ···················· |
| D | 8422 | hCoV-19/England/OXON-B19F4/2020\|EPI_ISL_479061\|202 | ···················· | ···················· | ····················· | ···················· | ···················· | ···················· |
| D | 8423 | hCoV-19/England/OXON-B1A00/2020\|EPI_ISL_479062\|202 | ···················· | ···················· | ····················· | ···················· | ···················· | ···················· |
| D | 8424 | hCoV-19/England/OXON-B1A1F/2020\|EPI_ISL_479063\|202 | ···················· | ···················· | ····················· | ···················· | ···················· | ···················· |
| D | 8425 | hCoV-19/England/OXON-B1A2E/2020\|EPI_ISL_479064\|202 | ···················· | ···················· | ····················· | ···················· | ···················· | ···················· |
| D | 8426 | hCoV-19/England/OXON-B1A4C/2020\|EPI_ISL_479066\|202 | ···················· | ·A·················· | ····················· | ···················· | ···················· | ···················· |
| D | 8427 | hCoV-19/England/OXON-B1A5B/2020\|EPI_ISL_479067\|202 | ···················· | ···················· | ····················· | ···················· | ···················· | ···················· |
| D | 8428 | hCoV-19/England/OXON-B1A79/2020\|EPI_ISL_479068\|202 | ···················· | ···················· | ····················· | ···················· | ···················· | ···················· |
| D | 8429 | hCoV-19/England/OXON-B1A88/2020\|EPI_ISL_479069\|202 | ···················· | ···················· | ····················· | ···················· | ···················· | ···················· |
| D | 8430 | hCoV-19/England/OXON-B1C28/2020\|EPI_ISL_479080\|202 | ···················· | ·A·················· | ····················· | ···················· | ···················· | ···················· |
| D | 8431 | hCoV-19/England/OXON-B1C55/2020\|EPI_ISL_479081\|202 | ···················· | ···················· | ····················· | ···················· | ···················· | ···················· |
| D | 8432 | hCoV-19/England/OXON-B1C64/2020\|EPI_ISL_479082\|202 | ···················· | ···················· | ····················· | ···················· | ···················· | ···················· |
| D | 8433 | hCoV-19/England/OXON-B1C91/2020\|EPI_ISL_479083\|202 | ···················· | ···················· | ····················· | ···················· | ···················· | ···················· |
| D | 8434 | hCoV-19/England/OXON-B1CBF/2020\|EPI_ISL_479084\|202 | ···················· | ···················· | ····················· | ···················· | ···················· | ···················· |
| D | 8435 | hCoV-19/England/OXON-B1CCE/2020\|EPI_ISL_479085\|202 | ···················· | ·A·················· | ····················· | ···················· | ···················· | ···················· |

| | | | | | | | | |
|---|---|---|---|---|---|---|---|---|
| D | 8436 | hCoV-19/England/OXON-B1CDD/2020\|EPI_ISL_479086\|202 | ···················· | ···················· | ···················· | ···················· | ···················· | ···················· |
| D | 8437 | hCoV-19/England/OXON-B1CFB/2020\|EPI_ISL_479088\|202 | ···················· | ···················· | ···················· | ···················· | ···················· | ···················· |
| D | 8438 | hCoV-19/England/OXON-B1D52/2020\|EPI_ISL_479089\|202 | ···················· | ·A·················· | ···················· | ···················· | ···················· | ···················· |
| D | 8439 | hCoV-19/England/OXON-B1D9E/2020\|EPI_ISL_479090\|202 | ···················· | ···················· | ···················· | ···················· | ···················· | ···················· |
| D | 8440 | hCoV-19/England/OXON-B1DAD/2020\|EPI_ISL_479091\|202 | ···················· | ···················· | ···················· | ···················· | ···················· | ···················· |
| D | 8441 | hCoV-19/England/OXON-B1DDA/2020\|EPI_ISL_479092\|202 | ···················· | ···················· | ···················· | ···················· | ···················· | ···················· |
| D | 8442 | hCoV-19/England/OXON-B1DE9/2020\|EPI_ISL_479093\|202 | ···················· | ···················· | ···················· | ···················· | ···················· | ···················· |
| D | 8443 | hCoV-19/England/OXON-B1DF8/2020\|EPI_ISL_479094\|202 | ···················· | ···················· | ···················· | ···················· | ···················· | ···················· |
| D | 8444 | hCoV-19/England/OXON-B1E22/2020\|EPI_ISL_479095\|202 | ···················· | ···················· | ···················· | ···················· | ···················· | ···················· |
| D | 8445 | hCoV-19/England/OXON-B1E31/2020\|EPI_ISL_479096\|202 | ···················· | ···················· | ···················· | ···················· | ···················· | ···················· |
| D | 8446 | hCoV-19/England/OXON-B1E6E/2020\|EPI_ISL_479097\|202 | ···················· | ···················· | ···················· | ···················· | ···················· | ···················· |
| D | 8447 | hCoV-19/England/OXON-B1E7D/2020\|EPI_ISL_479098\|202 | ···················· | ···················· | ···················· | ···················· | ···················· | ···················· |
| D | 8448 | hCoV-19/England/OXON-B1EAA/2020\|EPI_ISL_479099\|202 | ···················· | ·A·················· | ···················· | ···················· | ···················· | ···················· |
| D | 8449 | hCoV-19/Canada/BC_24228539/2020\|EPI_ISL_463200\|202 | ···················· | ·············A······ | ···················· | ···················· | ···················· | ···················· |
| D | 8450 | hCoV-19/Canada/BC_26624482/2020\|EPI_ISL_463245\|202 | ···················· | ···················· | ···················· | ···················· | ···················· | ···················· |
| D | 8451 | hCoV-19/Canada/BC_26625053/2020\|EPI_ISL_463246\|202 | ···················· | ···················· | ···················· | ···················· | ···················· | ···················· |
| D | 8452 | hCoV-19/Canada/BC_26628654/2020\|EPI_ISL_463247\|202 | ···················· | ···················· | ···················· | ···················· | ···················· | ···················· |
| D | 8453 | hCoV-19/Canada/BC_26646523/2020\|EPI_ISL_463248\|202 | ···················· | ···················· | ···················· | ···················· | ···················· | ···················· |
| D | 8454 | hCoV-19/Canada/BC_26649386/2020\|EPI_ISL_463249\|202 | ···················· | ···················· | ···················· | ···················· | ···················· | ···················· |
| D | 8455 | hCoV-19/Canada/BC_26650347/2020\|EPI_ISL_463250\|202 | ···················· | ···················· | ···················· | ···················· | ···················· | ···················· |
| D | 8456 | hCoV-19/Canada/BC_26654155/2020\|EPI_ISL_463251\|202 | ···················· | ···················· | ···················· | ···················· | ···················· | ···················· |
| D | 8457 | hCoV-19/Canada/BC_26656159/2020\|EPI_ISL_463252\|202 | ···················· | ···················· | ···················· | ···················· | ···················· | ···················· |
| D | 8458 | hCoV-19/Canada/BC_26659043/2020\|EPI_ISL_463253\|202 | ···················· | ···················· | ···················· | ···················· | ···················· | ···················· |
| D | 8459 | hCoV-19/Canada/BC_26693628/2020\|EPI_ISL_463254\|202 | ···················· | ···················· | ···················· | ···················· | ···················· | ···················· |
| D | 8460 | hCoV-19/Canada/BC_26943931/2020\|EPI_ISL_463255\|202 | ···················· | ···················· | ···················· | ···················· | ···················· | ···················· |
| D | 8461 | hCoV-19/Canada/BC_26964094/2020\|EPI_ISL_463256\|202 | ···················· | ···················· | ···················· | ···················· | ···················· | ···················· |
| D | 8462 | hCoV-19/Canada/BC_27007571/2020\|EPI_ISL_463257\|202 | ···················· | ···················· | ···················· | ···················· | ···················· | ···················· |
| D | 8463 | hCoV-19/Canada/BC_27017413/2020\|EPI_ISL_463258\|202 | ···················· | ···················· | ···················· | ···················· | ···················· | ···················· |
| D | 8464 | hCoV-19/Canada/BC_27081621/2020\|EPI_ISL_463259\|202 | ···················· | ·············A······ | ···················· | ···················· | ···················· | ···················· |
| D | 8465 | hCoV-19/Canada/BC_27121841/2020\|EPI_ISL_463260\|202 | ···················· | ···················· | ···················· | ···················· | ···················· | ···················· |
| D | 8466 | hCoV-19/Canada/BC_27180562/2020\|EPI_ISL_463261\|202 | ···················· | ···················· | ···················· | ···················· | ···················· | ···················· |
| D | 8467 | hCoV-19/Canada/BC_27222254/2020\|EPI_ISL_463262\|202 | ···················· | ···················· | ···················· | ···················· | ···················· | ···················· |
| D | 8468 | hCoV-19/Canada/BC_27279444/2020\|EPI_ISL_463263\|202 | ···················· | ···················· | ···················· | ···················· | ···················· | ···················· |
| D | 8469 | hCoV-19/Canada/BC_27280425/2020\|EPI_ISL_463264\|202 | ···················· | ···················· | ···················· | ···················· | ···················· | ···················· |
| D | 8470 | hCoV-19/Canada/BC_27280641/2020\|EPI_ISL_463265\|202 | ···················· | ···················· | ···················· | ···················· | ···················· | ···················· |
| D | 8471 | hCoV-19/Canada/BC_27280776/2020\|EPI_ISL_463266\|202 | ···················· | ···················· | ···················· | ···················· | ···················· | ···················· |
| D | 8472 | hCoV-19/Canada/BC_27283916/2020\|EPI_ISL_463267\|202 | ···················· | ···················· | ···················· | ···················· | ···················· | ···················· |
| D | 8473 | hCoV-19/Canada/BC_27285211/2020\|EPI_ISL_463268\|202 | ···················· | ···················· | ···················· | ···················· | ···················· | ···················· |
| D | 8474 | hCoV-19/Canada/BC_27298990/2020\|EPI_ISL_463270\|202 | ···················· | ···················· | ···················· | ···················· | ···················· | ···················· |
| D | 8475 | hCoV-19/Canada/BC_27302671/2020\|EPI_ISL_463271\|202 | ···················· | ···················· | ···················· | ···················· | ···················· | ···················· |
| D | 8476 | hCoV-19/Canada/BC_27303787/2020\|EPI_ISL_463272\|202 | ···················· | ···················· | ···················· | ···················· | ···················· | ···················· |
| D | 8477 | hCoV-19/Canada/BC_27303944/2020\|EPI_ISL_463273\|202 | ···················· | ···················· | ···················· | ···················· | ···················· | ···················· |
| D | 8478 | hCoV-19/Canada/BC_27304064/2020\|EPI_ISL_463274\|202 | ···················· | ···················· | ···················· | ···················· | ···················· | ···················· |
| D | 8479 | hCoV-19/Canada/BC_27304554/2020\|EPI_ISL_463275\|202 | ···················· | ···················· | ···················· | ···················· | ···················· | ···················· |
| D | 8480 | hCoV-19/Canada/BC_27305318/2020\|EPI_ISL_463276\|202 | ···················· | ···················· | ···················· | ···················· | ···················· | ···················· |
| D | 8481 | hCoV-19/USA/LA-BIE-029/2020\|EPI_ISL_463277\|2020-04 | ···················· | ···················· | ···················· | ···················· | ···················· | ···················· |
| D | 8482 | hCoV-19/USA/LA-BIE-030/2020\|EPI_ISL_463278\|2020-04 | ···················· | ···················· | ···················· | ···················· | ···················· | ···················· |
| D | 8483 | hCoV-19/USA/LA-BIE-031/2020\|EPI_ISL_463279\|2020-04 | ···················· | ···················· | ···················· | ···················· | ···················· | ···················· |
| D | 8484 | hCoV-19/USA/LA-BIE-032/2020\|EPI_ISL_463280\|2020-04 | ···················· | ···················· | ···················· | ···················· | ···················· | ···················· |
| D | 8485 | hCoV-19/USA/LA-BIE-033/2020\|EPI_ISL_463281\|2020-04 | ···················· | ···················· | ···················· | ···················· | ···················· | ···················· |
| D | 8486 | hCoV-19/USA/LA-BIE-034/2020\|EPI_ISL_463282\|2020-04 | ···················· | ···················· | ···················· | ···················· | ···················· | ···················· |
| D | 8487 | hCoV-19/USA/LA-BIE-035/2020\|EPI_ISL_463283\|2020-04 | ···················· | ···················· | ···················· | ···················· | ···················· | ···················· |
| D | 8488 | hCoV-19/USA/LA-BIE-036/2020\|EPI_ISL_463284\|2020-04 | ···················· | ···················· | ···················· | ···················· | ···················· | ···················· |
| D | 8489 | hCoV-19/USA/LA-BIE-037/2020\|EPI_ISL_463285\|2020-04 | ···················· | ···················· | ···················· | ···················· | ···················· | ···················· |
| D | 8490 | hCoV-19/USA/LA-BIE-038/2020\|EPI_ISL_463286\|2020-04 | ···················· | ···················· | ···················· | ···················· | ···················· | ···················· |
| D | 8491 | hCoV-19/USA/LA-BIE-039/2020\|EPI_ISL_463287\|2020-04 | ···················· | ···················· | ···················· | ···················· | ···················· | ···················· |
| D | 8492 | hCoV-19/USA/LA-BIE-040/2020\|EPI_ISL_463288\|2020-04 | ···················· | ···················· | ···················· | ···················· | ···················· | ···················· |
| D | 8493 | hCoV-19/USA/LA-BIE-041/2020\|EPI_ISL_463289\|2020-04 | ···················· | ···················· | ···················· | ···················· | ···················· | ···················· |
| D | 8494 | hCoV-19/USA/LA-BIE-042/2020\|EPI_ISL_463290\|2020-04 | ···················· | ···················· | ···················· | ···················· | ···················· | ···················· |
| D | 8495 | hCoV-19/USA/LA-BIE-043/2020\|EPI_ISL_463291\|2020-04 | ···················· | ···················· | ···················· | ···················· | ···················· | ···················· |
| D | 8496 | hCoV-19/USA/LA-BIE-044/2020\|EPI_ISL_463292\|2020-04 | ···················· | ···················· | ···················· | ···················· | ···················· | ···················· |
| D | 8497 | hCoV-19/USA/LA-BIE-045/2020\|EPI_ISL_463293\|2020-04 | ···················· | ···················· | ···················· | ···················· | ···················· | ···················· |
| D | 8498 | hCoV-19/USA/LA-BIE-046/2020\|EPI_ISL_463294\|2020-04 | ···················· | ···················· | ···················· | ···················· | ···················· | ···················· |
| D | 8499 | hCoV-19/USA/LA-BIE-047/2020\|EPI_ISL_463295\|2020-04 | ···················· | ···················· | ···················· | ···················· | ···················· | ···················· |
| D | 8500 | hCoV-19/USA/LA-BIE-048/2020\|EPI_ISL_463296\|2020-04 | ···················· | ···················· | ···················· | ···················· | ···················· | ···················· |
| D | 8501 | hCoV-19/USA/LA-BIE-049/2020\|EPI_ISL_463297\|2020-04 | ···················· | ···················· | ···················· | ···················· | ···················· | ···················· |
| D | 8502 | hCoV-19/USA/LA-BIE-050/2020\|EPI_ISL_463298\|2020-04 | ···················· | ···················· | ···················· | ···················· | ···················· | ···················· |
| D | 8503 | hCoV-19/USA/LA-BIE-091/2020\|EPI_ISL_463299\|2020-04 | ···················· | ···················· | ···················· | ···················· | ····T··············· | ···················· |
| D | 8504 | hCoV-19/England/20109007104/2020\|EPI_ISL_464525\|20 | ···················· | ···················· | ···················· | ···················· | ···················· | ···················· |
| D | 8505 | hCoV-19/England/20109007304/2020\|EPI_ISL_464527\|20 | ···················· | ···················· | ···················· | ···················· | ···················· | ···················· |
| D | 8506 | hCoV-19/England/20109007404/2020\|EPI_ISL_464528\|20 | ···················· | ···················· | ···················· | ···················· | ···················· | ···················· |
| D | 8507 | hCoV-19/England/20109007504/2020\|EPI_ISL_464529\|20 | ···················· | ···················· | ···················· | ···················· | ···················· | ···················· |
| D | 8509 | hCoV-19/England/201090677/2020\|EPI_ISL_464595\|2020 | ···················· | ···················· | ···················· | ···················· | ···················· | ···················· |
| D | 8510 | hCoV-19/England/20139064704/2020\|EPI_ISL_465854\|20 | ···················· | ···················· | ···················· | ···················· | ···················· | ···················· |
| D | 8511 | hCoV-19/England/20139066504/2020\|EPI_ISL_465855\|20 | ···················· | ···················· | ···················· | ···················· | ···················· | ···················· |
| D | 8512 | hCoV-19/England/20139066604/2020\|EPI_ISL_465856\|20 | ···················· | ···················· | ···················· | ···················· | ···················· | ···················· |
| D | 8513 | hCoV-19/England/20139067404/2020\|EPI_ISL_465858\|20 | ···················· | ···················· | ···················· | ···················· | ···················· | ···················· |
| D | 8514 | hCoV-19/England/20139069604/2020\|EPI_ISL_465859\|20 | ···················· | ···················· | ···················· | ···················· | ···················· | ···················· |

| | | | | | | | | |
|---|---|---|---|---|---|---|---|---|
| D | 8515 | hCoV-19/England/20139069704/2020\|EPI_ISL_465860\|20 | ···················· | ···················· | ····················· | ···················· | ···················· | ···················· |
| D | 8516 | hCoV-19/England/20139069804/2020\|EPI_ISL_465861\|20 | ···················· | ···················· | ····················· | ···················· | ···················· | ···················· |
| D | 8519 | hCoV-19/Switzerland/ZH-ETHZ-130026/2020\|EPI_ISL_46 | ···················· | ···················· | ····················· | ···················· | ···················· | ···················· |
| D | 8520 | hCoV-19/India/GJ-GBRC169a/2020\|EPI_ISL_467029\|2020 | ···················· | ···················· | ····················· | ···················· | ···················· | ···················· |
| D | 8521 | hCoV-19/India/GJ-GBRC169b/2020\|EPI_ISL_467030\|2020 | ···················· | ·A·················· | ····················· | ···················· | ···················· | ···················· |
| D | 8522 | hCoV-19/India/GJ-GBRC170a/2020\|EPI_ISL_467031\|2020 | ···················· | ···················· | ····················· | ···················· | ···················· | ···················· |
| D | 8523 | hCoV-19/India/GJ-GBRC170b/2020\|EPI_ISL_467032\|2020 | ···················· | ·A·················· | ····················· | ···················· | ···················· | ···················· |
| D | 8524 | hCoV-19/India/GJ-GBRC171/2020\|EPI_ISL_467033\|2020- | ···················· | ···················· | ····················· | ···················· | ···················· | ···················· |
| D | 8525 | hCoV-19/India/GJ-GBRC172/2020\|EPI_ISL_467034\|2020- | ···················· | ·A·················· | ····················· | ···················· | ···················· | ···A················ |
| D | 8526 | hCoV-19/India/GJ-GBRC173a/2020\|EPI_ISL_467035\|2020 | ···················· | ···················· | ····················· | ···················· | ···················· | ···················· |
| D | 8527 | hCoV-19/India/GJ-GBRC173b/2020\|EPI_ISL_467036\|2020 | ···················· | ···················· | ····················· | ···················· | ···················· | ···················· |
| D | 8528 | hCoV-19/India/GJ-GBRC174a/2020\|EPI_ISL_467037\|2020 | ···················· | ···················· | ····················· | ···················· | ···················· | ···················· |
| D | 8529 | hCoV-19/India/GJ-GBRC174b/2020\|EPI_ISL_467038\|2020 | ···················· | ·A·················· | ····················· | ···················· | ···················· | ···A················ |
| D | 8530 | hCoV-19/India/GJ-GBRC175/2020\|EPI_ISL_467039\|2020- | ···················· | ···················· | ····················· | ···················· | ···················· | ···················· |
| D | 8531 | hCoV-19/India/GJ-GBRC176/2020\|EPI_ISL_467040\|2020- | ···················· | ···················· | ····················· | ···················· | ···················· | ···················· |
| D | 8532 | hCoV-19/India/GJ-GBRC177a/2020\|EPI_ISL_467041\|2020 | ···················· | ·A·················· | ····················· | ···················· | ···················· | ···A················ |
| D | 8533 | hCoV-19/India/GJ-GBRC177b/2020\|EPI_ISL_467042\|2020 | ···················· | ···················· | ····················· | ···················· | ···················· | ···A················ |
| D | 8534 | hCoV-19/India/GJ-GBRC178a/2020\|EPI_ISL_467043\|2020 | ···················· | ···················· | ····················· | ···················· | ···················· | ···················· |
| D | 8535 | hCoV-19/India/GJ-GBRC178b/2020\|EPI_ISL_467044\|2020 | ···················· | ·A·················· | ····················· | ···················· | ···················· | ···················· |
| D | 8536 | hCoV-19/India/GJ-GBRC179a/2020\|EPI_ISL_467045\|2020 | ···················· | ·A·················· | ····················· | ···················· | ···················· | ···A················ |
| D | 8537 | hCoV-19/India/GJ-GBRC179b/2020\|EPI_ISL_467046\|2020 | ···················· | ···················· | ····················· | ···················· | ···················· | ···A················ |
| D | 8538 | hCoV-19/India/GJ-GBRC180a/2020\|EPI_ISL_467047\|2020 | ···················· | ···················· | ····················· | ···················· | ···················· | ···················· |
| D | 8539 | hCoV-19/India/GJ-GBRC180b/2020\|EPI_ISL_467048\|2020 | ···················· | ·A·················· | ····················· | ···················· | ···················· | ···················· |
| D | 8540 | hCoV-19/India/GJ-GBRC181a/2020\|EPI_ISL_467049\|2020 | ···················· | ·A·················· | ····················· | ···················· | ···················· | ···A················ |
| D | 8541 | hCoV-19/Spain/GA-USC-009/2020\|EPI_ISL_468339\|2020- | ···················· | ···················· | ····················· | ···················· | ···················· | ···················· |
| D | 8542 | hCoV-19/Spain/GA-USC-010/2020\|EPI_ISL_468340\|2020- | ···················· | ···················· | ····················· | ···················· | ···················· | ···················· |
| D | 8543 | hCoV-19/USA/CA-CZB-1284/2020\|EPI_ISL_468345\|2020-0 | ···················· | ···················· | ····················· | ···················· | ···················· | ···················· |
| D | 8544 | hCoV-19/USA/CA-CZB-1286/2020\|EPI_ISL_468346\|2020-0 | ···················· | ···················· | ····················· | ···················· | ···················· | ···················· |
| D | 8545 | hCoV-19/USA/CA-CZB-1291/2020\|EPI_ISL_468347\|2020-0 | ···················· | ···················· | ····················· | ···················· | ···················· | ···················· |
| D | 8546 | hCoV-19/USA/CA-CZB-1295/2020\|EPI_ISL_468348\|2020-0 | ···················· | ···················· | ····················· | ···················· | ···················· | ···················· |
| D | 8547 | hCoV-19/USA/CA-CZB-1304/2020\|EPI_ISL_468349\|2020-0 | ···················· | ···················· | ····················· | ···················· | ···················· | ···················· |
| D | 8548 | hCoV-19/USA/CA-CZB-1309/2020\|EPI_ISL_468350\|2020-0 | ···················· | ···················· | ····················· | ···················· | ···················· | ···················· |
| D | 8549 | hCoV-19/USA/CA-CZB-1310/2020\|EPI_ISL_468351\|2020-0 | ···················· | ···················· | ····················· | ···················· | ···················· | ···················· |
| D | 8550 | hCoV-19/USA/CA-CZB-1317/2020\|EPI_ISL_468353\|2020-0 | ···················· | ···················· | ····················· | ···················· | ···················· | ···················· |
| D | 8551 | hCoV-19/USA/CA-CZB-1320/2020\|EPI_ISL_468354\|2020-0 | ···················· | ···················· | ····················· | ···················· | ···················· | ···················· |
| D | 8552 | hCoV-19/USA/CA-CZB-1322/2020\|EPI_ISL_468355\|2020-0 | ···················· | ···················· | ····················· | ···················· | ···················· | ···················· |
| D | 8553 | hCoV-19/USA/CA-CZB-1324/2020\|EPI_ISL_468356\|2020-0 | ···················· | ···················· | ····················· | ···················· | ···················· | ···················· |
| D | 8554 | hCoV-19/USA/CA-CZB-1330/2020\|EPI_ISL_468358\|2020-0 | ···················· | ···················· | ····················· | ···················· | ···················· | ···················· |
| D | 8555 | hCoV-19/USA/CA-CZB-1341/2020\|EPI_ISL_468360\|2020-0 | ···················· | ···················· | ···········T········· | ···················· | ···················· | ···················· |
| D | 8556 | hCoV-19/USA/CA-CZB-1343/2020\|EPI_ISL_468361\|2020-0 | ···················· | ···················· | ····················· | ···················· | ···················· | ···················· |
| D | 8557 | hCoV-19/USA/CA-CZB-1347/2020\|EPI_ISL_468363\|2020-0 | ···················· | ···················· | ····················· | ···················· | ···················· | ···················· |
| D | 8558 | hCoV-19/USA/CA-CZB-1348/2020\|EPI_ISL_468364\|2020-0 | ···················· | ···················· | ····················· | ···················· | ···················· | ···················· |
| D | 8559 | hCoV-19/USA/CA-CZB-1349/2020\|EPI_ISL_468365\|2020-0 | ···················· | ···················· | ····················· | ···················· | ···················· | ···················· |
| D | 8560 | hCoV-19/USA/CA-CZB-1353/2020\|EPI_ISL_468367\|2020-0 | ···················· | ···················· | ····················· | ···················· | ···················· | ···················· |
| D | 8561 | hCoV-19/USA/CA-CZB-1354/2020\|EPI_ISL_468368\|2020-0 | ···················· | ···················· | ····················· | ···················· | ···················· | ···················· |
| D | 8562 | hCoV-19/USA/CA-CZB-1356/2020\|EPI_ISL_468370\|2020-0 | ···················· | ···················· | ····················· | ···················· | ···················· | ···················· |
| D | 8563 | hCoV-19/USA/CA-CZB-1360/2020\|EPI_ISL_468372\|2020-0 | ···················· | ···················· | ····················· | ···················· | ···················· | ···················· |
| D | 8564 | hCoV-19/USA/CA-CZB-1364/2020\|EPI_ISL_468373\|2020-0 | ···················· | ···················· | ····················· | ···················· | ···················· | ···················· |
| D | 8565 | hCoV-19/USA/CA-CZB-1369/2020\|EPI_ISL_468374\|2020-0 | ···················· | ···················· | ····················· | ···················· | ···················· | ···················· |
| D | 8566 | hCoV-19/USA/CA-CZB-1393/2020\|EPI_ISL_468377\|2020-0 | ···················· | ···················· | ····················· | ···················· | ···················· | ···················· |
| D | 8567 | hCoV-19/England/BRIS-127EAE/2020\|EPI_ISL_469625\|20 | ···················· | ···················· | ····················· | ···················· | ···················· | ···················· |
| D | 8568 | hCoV-19/England/BRIS-126EFA/2020\|EPI_ISL_469626\|20 | ···················· | ····A··············· | ····················· | ···················· | ···················· | ···················· |
| D | 8569 | hCoV-19/England/BRIS-127AD7/2020\|EPI_ISL_469629\|20 | ···················· | ···················· | ····················· | ···················· | ···················· | ···················· |
| D | 8570 | hCoV-19/England/BRIS-1278CE/2020\|EPI_ISL_469670\|20 | ···················· | ···················· | ····················· | ···················· | ···················· | ···················· |
| D | 8571 | hCoV-19/England/BRIS-127BF2/2020\|EPI_ISL_469674\|20 | ···················· | ···················· | ····················· | ···················· | ···················· | ···················· |
| D | 8572 | hCoV-19/England/BRIS-1279CB/2020\|EPI_ISL_469675\|20 | ···················· | ···················· | ····················· | ···················· | ···················· | ···················· |
| D | 8573 | hCoV-19/England/BRIS-1284E7/2020\|EPI_ISL_469676\|20 | ···················· | ···················· | ····················· | ···················· | ···················· | ···················· |
| D | 8574 | hCoV-19/England/BRIS-12865A/2020\|EPI_ISL_469677\|20 | ···················· | ···················· | ····················· | ···················· | ···················· | ···················· |
| D | 8575 | hCoV-19/England/BRIS-127451/2020\|EPI_ISL_469679\|20 | ···················· | ···················· | ····················· | ···················· | ···················· | ···················· |
| D | 8576 | hCoV-19/England/BRIS-128292/2020\|EPI_ISL_469680\|20 | ···················· | ···················· | ····················· | ···················· | ···················· | ···················· |
| D | 8577 | hCoV-19/England/BRIS-1278A0/2020\|EPI_ISL_469681\|20 | ···················· | ···················· | ····················· | ···················· | ···················· | ···················· |
| D | 8578 | hCoV-19/England/BRIS-127E80/2020\|EPI_ISL_469682\|20 | ···················· | ···················· | ····················· | ···················· | ···················· | ···················· |
| D | 8579 | hCoV-19/England/BRIS-1283BD/2020\|EPI_ISL_469683\|20 | ···················· | ···················· | ····················· | ···················· | ···················· | ···················· |
| D | 8580 | hCoV-19/England/BRIS-12712D/2020\|EPI_ISL_469684\|20 | ···················· | ···················· | ····················· | ···················· | ···················· | ···················· |
| D | 8581 | hCoV-19/England/BRIS-1272C0/2020\|EPI_ISL_469686\|20 | ···················· | ···················· | ····················· | ···················· | ···················· | ···················· |
| D | 8582 | hCoV-19/England/BRIS-127E53/2020\|EPI_ISL_469687\|20 | ···················· | ···················· | ····················· | ···················· | ···················· | ···················· |
| D | 8583 | hCoV-19/England/BRIS-127BA7/2020\|EPI_ISL_469691\|20 | ···················· | ····A··············· | ····················· | ···················· | ···················· | ···················· |
| D | 8584 | hCoV-19/England/BRIS-1273BE/2020\|EPI_ISL_469692\|20 | ···················· | ···················· | ····················· | ···················· | ···················· | ···················· |
| D | 8585 | hCoV-19/England/BRIS-127952/2020\|EPI_ISL_469693\|20 | ···················· | ···················· | ····················· | ···················· | ···················· | ···················· |
| D | 8586 | hCoV-19/England/BRIS-128414/2020\|EPI_ISL_469694\|20 | ···················· | ···················· | ····················· | ···················· | ···················· | ···················· |
| D | 8587 | hCoV-19/England/BRIS-12749D/2020\|EPI_ISL_469696\|20 | ···················· | ···················· | ····················· | ···················· | ···················· | ···················· |
| D | 8588 | hCoV-19/England/BRIS-12707B/2020\|EPI_ISL_469697\|20 | ···················· | ···················· | ····················· | ···················· | ···················· | ···················· |
| D | 8589 | hCoV-19/USA/UT-01867/2020\|EPI_ISL_470691\|2020-04-1 | ···················· | ···················· | ····················· | ···················· | ···················· | ···················· |
| D | 8590 | hCoV-19/England/EXET-136FB8/2020\|EPI_ISL_471941\|20 | ···················· | ···················· | ····················· | ···················· | ···················· | ···················· |
| D | 8591 | hCoV-19/England/EXET-136FE5/2020\|EPI_ISL_471943\|20 | ···················· | ···················· | ····················· | ···················· | ···················· | ···················· |
| D | 8592 | hCoV-19/England/EXET-136FF4/2020\|EPI_ISL_471944\|20 | ···················· | ·A·················· | ····················· | ···················· | ···················· | ···················· |
| D | 8593 | hCoV-19/England/EXET-13705A/2020\|EPI_ISL_471946\|20 | ···················· | ···················· | ····················· | ···················· | ···················· | ···················· |
| D | 8594 | hCoV-19/England/EXET-137069/2020\|EPI_ISL_471947\|20 | ···················· | ···················· | ····················· | ···················· | ···················· | ···················· |

D 8595 hCoV-19/England/EXET-1370F0/2020|EPI_ISL_471949|20 ···················· ···················· ···················· ···················· ···················· ····················
D 8596 hCoV-19/England/EXET-13710C/2020|EPI_ISL_471950|20 ···················· ···················· ···················· ···················· ···················· ····················
D 8599 hCoV-19/England/EXET-137236/2020|EPI_ISL_471956|20 ···················· ···················· ···················· ···················· ···················· ····················
D 8600 hCoV-19/England/EXET-137245/2020|EPI_ISL_471957|20 ···················· ···················· ···················· ···················· ···················· ····················
D 8601 hCoV-19/England/EXET-1372CD/2020|EPI_ISL_471959|20 ···················· ···················· ···················· ···················· ···················· ····················
D 8602 hCoV-19/England/EXET-1372FA/2020|EPI_ISL_471960|20 ···················· ···················· ···················· ···················· ···················· ····················
D 8604 hCoV-19/England/EXET-137342/2020|EPI_ISL_471964|20 ···················· ···················· ···················· ···················· ···················· ····················
D 8605 hCoV-19/England/EXET-137360/2020|EPI_ISL_471966|20 ···················· ·············A······ ···················· ···················· ···················· ····················
D 8606 hCoV-19/England/EXET-13737F/2020|EPI_ISL_471967|20 ···················· ···················· ···················· ···················· ···················· ····················
D 8609 hCoV-19/England/EXET-1373E8/2020|EPI_ISL_471973|20 ···················· ···················· ···················· ···················· ···················· ····················
D 8610 hCoV-19/England/EXET-137403/2020|EPI_ISL_471975|20 ···················· ···················· ···················· ···················· ···················· ····················
D 8613 hCoV-19/England/EXET-1375C4/2020|EPI_ISL_471978|20 ···················· ···················· ···················· ···················· ···················· ····················
D 8614 hCoV-19/England/EXET-1375F1/2020|EPI_ISL_471979|20 ···················· ···················· ···················· ···················· ···················· ····················
D 8615 hCoV-19/USA/NE-11518/2020|EPI_ISL_475225|2020-05-0 ···················· ···················· ···················· ···················· ···················· ····················
D 8616 hCoV-19/USA/NE-11527/2020|EPI_ISL_475227|2020-05-0 ···················· ···················· ···················· ···················· ···················· ····················
D 8617 hCoV-19/USA/NE-11549/2020|EPI_ISL_475228|2020-05-0 ···················· ···················· ···················· ···················· ···················· ····················
D 8619 hCoV-19/USA/NE-11566/2020|EPI_ISL_475231|2020-05-0 ···················· ···················· ···················· ···················· ···················· ····················
D 8620 hCoV-19/USA/CA-11567/2020|EPI_ISL_475232|2020-05-0 ···················· ··C················· ···················· ···················· ···················· ····················
D 8621 hCoV-19/USA/NE-11644/2020|EPI_ISL_475233|2020-05-0 ···················· ···················· ···················· ···················· ···················· ····················
D 8622 hCoV-19/USA/NE-11837/2020|EPI_ISL_475236|2020-05-0 ···················· ···················· ···················· ···················· ···················· ····················
D 8623 hCoV-19/USA/NE-11845/2020|EPI_ISL_475237|2020-05-0 ···················· ···················· ···················· ···················· ···················· ····················
D 8624 hCoV-19/Bangladesh/BCSIR-NILMRC-114/2020|EPI_ISL_4 ···················· ···················· ···················· ···················· ···················· ····················
D 8625 hCoV-19/England/PORT-2CF8C7/2020|EPI_ISL_475240|20 ···················· ···················· ···················· ···················· ···················· ····················
D 8626 hCoV-19/England/PORT-2CFA76/2020|EPI_ISL_475241|20 ···················· ···················· ···················· ···················· ···················· ····················
D 8627 hCoV-19/England/PORT-2CFE7A/2020|EPI_ISL_475242|20 ···················· ···················· ···················· ···················· ···················· ····················
D 8628 hCoV-19/England/PORT-2CFEB6/2020|EPI_ISL_475244|20 ···················· ···················· ···················· ···················· ···················· ····················
D 8629 hCoV-19/England/PORT-2CFEF2/2020|EPI_ISL_475247|20 ···················· ···················· ···················· ···················· ···················· ····················
D 8631 hCoV-19/England/PORT-2D0C30/2020|EPI_ISL_475266|20 ···················· ···················· ···················· ···················· ···················· ····················
D 8632 hCoV-19/England/PORT-2D0C4F/2020|EPI_ISL_475267|20 ···················· ···················· ···················· ···················· ···················· ····················
D 8633 hCoV-19/England/PORT-2D0C7C/2020|EPI_ISL_475268|20 ···················· ···················· ···················· ···················· ···················· ····················
D 8634 hCoV-19/England/PORT-2D0C8B/2020|EPI_ISL_475269|20 ···················· ···················· ···················· ···················· ···················· ····················
D 8635 hCoV-19/England/PORT-2D0CA9/2020|EPI_ISL_475271|20 ···················· ···················· ···················· ···················· ···················· ····················
D 8636 hCoV-19/England/PORT-2D0CB8/2020|EPI_ISL_475272|20 ···················· ···················· ···················· ···················· ···················· ····················
D 8637 hCoV-19/England/PORT-2D0CD6/2020|EPI_ISL_475273|20 ···················· ···················· ···················· ···················· ···················· ····················
D 8638 hCoV-19/England/PORT-2D0CE5/2020|EPI_ISL_475274|20 ···················· ···················· ···················· ···················· ···················· ····················
D 8639 hCoV-19/England/PORT-2D0D2E/2020|EPI_ISL_475276|20 ···················· ···················· ···················· ···················· ···················· ····················
D 8640 hCoV-19/England/PORT-2D0D6A/2020|EPI_ISL_475279|20 ···················· ···················· ···················· ···················· ···················· ····················
D 8641 hCoV-19/England/PORT-2D0D79/2020|EPI_ISL_475280|20 ···················· ···················· ···················· ···················· ···················· ····················
D 8642 hCoV-19/England/PORT-2D0D97/2020|EPI_ISL_475282|20 ···················· ···················· ···················· ···················· ···················· ····················
D 8643 hCoV-19/England/PORT-2D0DA6/2020|EPI_ISL_475283|20 ···················· ···················· ···················· ···················· ···················· ····················
D 8644 hCoV-19/England/PORT-2D0DB5/2020|EPI_ISL_475284|20 ···················· ···················· ···················· ···················· ···················· ····················
D 8645 hCoV-19/England/PORT-2D0DC4/2020|EPI_ISL_475285|20 ···················· ···················· ···················· ···················· ···················· ····················
D 8646 hCoV-19/England/PORT-2D0DD3/2020|EPI_ISL_475286|20 ···················· ···················· ···················· ···················· ···················· ····················
D 8647 hCoV-19/England/PORT-2D0DE2/2020|EPI_ISL_475287|20 ···················· ···················· ···················· ···················· ···················· ····················
D 8648 hCoV-19/England/PORT-2D0DF1/2020|EPI_ISL_475288|20 ···················· ···················· ···················· ···················· ···················· ····················
D 8649 hCoV-19/England/PORT-2D0E0D/2020|EPI_ISL_475289|20 ···················· ···················· ···················· ···················· ···················· ····················
D 8650 hCoV-19/England/PORT-2D0E2B/2020|EPI_ISL_475291|20 ···················· ···················· ···················· ···················· ···················· ····················
D 8651 hCoV-19/England/PORT-2D0E3A/2020|EPI_ISL_475292|20 ···················· ···················· ···················· ···················· ···················· ····················
D 8652 hCoV-19/England/PORT-2D0E49/2020|EPI_ISL_475293|20 ···················· ···················· ···················· ···················· ···················· ····················
D 8653 hCoV-19/England/PORT-2D0E76/2020|EPI_ISL_475296|20 ···················· ···················· ···················· ···················· ···················· ····················
D 8654 hCoV-19/England/PORT-2D0E85/2020|EPI_ISL_475297|20 ···················· ···················· ···················· ···················· ···················· ····················
D 8655 hCoV-19/England/PORT-2D0E94/2020|EPI_ISL_475298|20 ···················· ···················· ···················· ···················· ···················· ····················
D 8656 hCoV-19/England/PORT-2D0EA3/2020|EPI_ISL_475299|20 ···················· ···················· ···················· ···················· ···················· ····················
D 8657 hCoV-19/Canada/QC_AQ3/2020|EPI_ISL_476511|2020-04- ···················· ···················· ···················· ···················· ···················· ····················
D 8658 hCoV-19/Canada/QC_AR2/2020|EPI_ISL_476512|2020-04- ···················· ···················· ···················· ···················· ···················· ····················
D 8659 hCoV-19/Canada/QC_AS5/2020|EPI_ISL_476513|2020-04- ···················· ···················· ···················· ···················· ···················· ····················
D 8660 hCoV-19/Senegal/63742/2020|EPI_ISL_476514|2020-06- ···················· ···················· ···················· ···················· ···················· ····················
D 8661 hCoV-19/Senegal/64101/2020|EPI_ISL_476516|2020-06- ···················· ···················· ···················· ···················· ···················· ····················
D 8662 hCoV-19/USA/CT-Yale-287/2020|EPI_ISL_476517|2020-0 ···················· ···················· ···················· ···················· ···················· ····················
D 8663 hCoV-19/USA/CT-Yale-288/2020|EPI_ISL_476518|2020-0 ···················· ···················· ···················· ···················· ···················· ····················
D 8664 hCoV-19/USA/CT-Yale-292/2020|EPI_ISL_476521|2020-0 ···················· ···················· ···················· ···················· ···················· ····················
D 8665 hCoV-19/USA/CT-Yale-297/2020|EPI_ISL_476526|2020-0 ···················· ···················· ···················· ···················· ···················· ····················
D 8666 hCoV-19/USA/CT-Yale-298/2020|EPI_ISL_476527|2020-0 ···················· ···················· ···················· ···················· ···················· ····················
D 8667 hCoV-19/USA/CT-Yale-299/2020|EPI_ISL_476528|2020-0 ···················· ···················· ···················· ···················· ···················· ····················
D 8668 hCoV-19/USA/CT-Yale-328/2020|EPI_ISL_476556|2020-0 ···················· ···················· ···················· ···················· ···················· ····················
D 8669 hCoV-19/Senegal/64166/2020|EPI_ISL_476558|2020-06- ···················· ···················· ···················· ···················· ···················· ····················
D 8670 hCoV-19/Morocco/refstage1/2020|EPI_ISL_476559|2020 ···················· ···················· ···················· ···················· ···················· ····················
D 8671 hCoV-19/Argentina/Heritas_HG006/2020|EPI_ISL_47656 ···················· ···················· ···················· ···················· ···················· ····················
D 8672 hCoV-19/Senegal/64763/2020|EPI_ISL_476562|2020-06- ···················· ···················· ···················· ···················· ···················· ····················
D 8673 hCoV-19/Argentina/Heritas_HG014/2020|EPI_ISL_47656 ···················· ···················· ···················· ···················· ···················· ····················
D 8674 hCoV-19/Senegal/64766/2020|EPI_ISL_476564|2020-06- ···················· ···················· ···················· ···················· ···················· ···············A····
D 8675 hCoV-19/Argentina/Heritas_HG007/2020|EPI_ISL_47656 ···················· ···················· ···················· ···················· ···················· ····················
D 8676 hCoV-19/Argentina/Heritas_HG015/2020|EPI_ISL_47656 ···················· ···················· ···················· ···················· ···················· ····················
D 8677 hCoV-19/Argentina/Heritas_HG009/2020|EPI_ISL_47656 ···················· ···················· ···················· ···················· ···················· ····················
D 8678 hCoV-19/Senegal/64932/2020|EPI_ISL_476569|2020-06- ···················· ···················· ···················· ···················· ···················· ····················
D 8679 hCoV-19/Argentina/Heritas_HG018/2020|EPI_ISL_47657 ···················· ···················· ···················· ···················· ···················· ····················
D 8680 hCoV-19/Senegal/65030/2020|EPI_ISL_476572|2020-06- ···················· ···················· ···················· ···················· ···················· ····················
D 8681 hCoV-19/Argentina/Heritas_HG019/2020|EPI_ISL_47657 ···················· ···················· ···················· ···················· ···················· ····················

| | | | | | | | | |
|---|---|---|---|---|---|---|---|---|
| D | 8682 hCoV-19/Senegal/65052/2020\|EPI_ISL_476574\|2020-06- | ···················· | ···················· | ····················· | ···················· | ···················· | ···················· | |
| D | 8683 hCoV-19/Scotland/CVR1986/2020\|EPI_ISL_477842\|2020- | ···················· | ···················· | ····················· | ···················· | ···················· | ···················· | |
| D | 8684 hCoV-19/Scotland/CVR2011/2020\|EPI_ISL_477843\|2020- | ···················· | ···················· | ····················· | ···················· | ···················· | ···················· | |
| D | 8685 hCoV-19/Scotland/CVR2036/2020\|EPI_ISL_477845\|2020- | ···················· | ···················· | ····················· | ···················· | ···················· | ···················· | |
| D | 8686 hCoV-19/Scotland/CVR2067/2020\|EPI_ISL_477849\|2020- | ···················· | ···················· | ····················· | ···················· | ···················· | ···················· | |
| D | 8687 hCoV-19/Scotland/CVR2086/2020\|EPI_ISL_477852\|2020- | ···················· | ···················· | ····················· | ···················· | ···················· | ···················· | |
| D | 8688 hCoV-19/Scotland/CVR2129/2020\|EPI_ISL_477857\|2020- | ···················· | ···················· | ····················· | ···················· | ···················· | ···················· | |
| D | 8689 hCoV-19/Scotland/CVR2130/2020\|EPI_ISL_477858\|2020- | ···················· | ···················· | ····················· | ···················· | ···················· | ···················· | |
| D | 8690 hCoV-19/Scotland/CVR2131/2020\|EPI_ISL_477859\|2020- | ···················· | ···················· | ····················· | ···················· | ···················· | ···················· | |
| D | 8691 hCoV-19/Scotland/CVR2255/2020\|EPI_ISL_477886\|2020- | ···················· | ·A·················· | ····················· | ···················· | ···················· | ···················· | |
| D | 8692 hCoV-19/Scotland/CVR2265/2020\|EPI_ISL_477887\|2020- | ···················· | ···················· | ····················· | ···················· | ···················· | ···················· | |
| D | 8693 hCoV-19/Scotland/CVR2271/2020\|EPI_ISL_477889\|2020- | ··T················· | ···················A | ····················· | ···················· | ···················· | ···················· | 1 |
| D | 8694 hCoV-19/Scotland/CVR2273/2020\|EPI_ISL_477890\|2020- | ···················· | ···················· | ····················· | ···················· | ···················· | ···················· | |
| D | 8695 hCoV-19/Scotland/CVR2288/2020\|EPI_ISL_477894\|2020- | ···················· | ···················· | ···········T········· | ···················· | ···················· | ···················· | |
| D | 8696 hCoV-19/Scotland/CVR2297/2020\|EPI_ISL_477895\|2020- | ···················· | ···················A | ····················· | ···················· | ···················· | ···················· | 1 |
| D | 8697 hCoV-19/Scotland/CVR2303/2020\|EPI_ISL_477896\|2020- | ···················· | ·A·················· | ····················· | ···················· | ···················· | ···················· | |
| D | 8698 hCoV-19/Scotland/CVR2305/2020\|EPI_ISL_477897\|2020- | ···················· | ···················· | ····················· | ···················· | ···················· | ···················· | |
| D | 8699 hCoV-19/Scotland/CVR2306/2020\|EPI_ISL_477898\|2020- | ···················· | ···················· | ····················· | ···················· | ···················· | ···················· | |
| D | 8700 hCoV-19/Scotland/CVR2307/2020\|EPI_ISL_477899\|2020- | ···················· | ···················· | ····················· | ···················· | ···················· | ···················· | |
| D | 8701 hCoV-19/England/OXON-B14C6/2020\|EPI_ISL_479020\|202 | ···················· | ···················· | ····················· | ···················· | ···················· | ···················· | |
| D | 8702 hCoV-19/England/OXON-B1596/2020\|EPI_ISL_479025\|202 | ···················· | ···················· | ····················· | ···················· | ···················· | ···················· | |
| D | 8703 hCoV-19/England/OXON-B15A5/2020\|EPI_ISL_479026\|202 | ···················· | ···················· | ····················· | ···················· | ···················· | ···················· | |
| D | 8704 hCoV-19/England/OXON-B15C3/2020\|EPI_ISL_479027\|202 | ···················· | ···················· | ····················· | ···················· | ···················· | ···················· | |
| D | 8705 hCoV-19/Canada/BC_24232310/2020\|EPI_ISL_463201\|202 | ···················· | ···················· | ····················· | ···················· | ···················· | ···················· | |
| D | 8706 hCoV-19/Canada/BC_24234949/2020\|EPI_ISL_463202\|202 | ···················· | ···················· | ····················· | ···················· | ···················· | ···················· | |
| D | 8707 hCoV-19/Canada/BC_24235415/2020\|EPI_ISL_463203\|202 | ···················· | ···················· | ····················· | ···················· | ···················· | ···················· | |
| D | 8708 hCoV-19/Canada/BC_24239917/2020\|EPI_ISL_463204\|202 | ···················· | ···················· | ····················· | ···················· | ···················· | ···················· | |
| D | 8709 hCoV-19/Canada/BC_24245374/2020\|EPI_ISL_463205\|202 | ···················· | ···················· | ····················· | ···················· | ···················· | ···················· | |
| D | 8710 hCoV-19/Canada/BC_24252365/2020\|EPI_ISL_463206\|202 | ···················· | ···················· | ····················· | ···················· | ···················· | ···················· | |
| D | 8711 hCoV-19/Canada/BC_24258528/2020\|EPI_ISL_463207\|202 | ···················· | ···················· | ····················· | ···················· | ···················· | ···················· | |
| D | 8712 hCoV-19/Canada/BC_24261010/2020\|EPI_ISL_463208\|202 | ···················· | ···················· | ····················· | ···················· | ···················· | ···················· | |
| D | 8713 hCoV-19/Canada/BC_24271871/2020\|EPI_ISL_463209\|202 | ···················· | ···················· | ····················· | ···················· | ···················· | ···················· | |
| D | 8714 hCoV-19/Canada/BC_24302962/2020\|EPI_ISL_463210\|202 | ···················· | ···················· | ····················· | ···················· | ···················· | ···················· | |
| D | 8715 hCoV-19/Canada/BC_24319756/2020\|EPI_ISL_463211\|202 | ···················· | ···················· | ····················· | ···················· | ···················· | ···················· | |
| D | 8716 hCoV-19/Canada/BC_24340030/2020\|EPI_ISL_463212\|202 | ···················· | ···················· | ····················· | ···················· | ···················· | ···················· | |
| D | 8717 hCoV-19/Canada/BC_24340319/2020\|EPI_ISL_463213\|202 | ···················· | ···················· | ····················· | ···················· | ···················· | ···················· | |
| D | 8718 hCoV-19/Canada/BC_24360831/2020\|EPI_ISL_463214\|202 | ···················· | ···················· | ····················· | ···················· | ···················· | ···················· | |
| D | 8719 hCoV-19/Canada/BC_24380644/2020\|EPI_ISL_463215\|202 | ···················· | ···················· | ····················· | ···················· | ···················· | ···················· | |
| D | 8720 hCoV-19/Canada/BC_24825751/2020\|EPI_ISL_463216\|202 | ···················· | ···················· | ····················· | ···················· | ···················· | ···················· | |
| D | 8721 hCoV-19/Canada/BC_25143276/2020\|EPI_ISL_463217\|202 | ···················· | ···················· | ····················· | ···················· | ···················· | ···················· | |
| D | 8722 hCoV-19/Canada/BC_25336669/2020\|EPI_ISL_463219\|202 | ···················· | ···················· | ····················· | ···················· | ···················· | ···················· | |
| D | 8723 hCoV-19/Canada/BC_25367465/2020\|EPI_ISL_463220\|202 | ···················· | ···················· | ····················· | ···················· | ···················· | ···················· | |
| D | 8724 hCoV-19/Canada/BC_25376719/2020\|EPI_ISL_463221\|202 | ···················· | ···················· | ····················· | ···················· | ···················· | ···················· | |
| D | 8725 hCoV-19/Canada/BC_25384316/2020\|EPI_ISL_463222\|202 | ···················· | ···················· | ····················· | ···················· | ···················· | ···················· | |
| D | 8726 hCoV-19/Canada/BC_25390851/2020\|EPI_ISL_463223\|202 | ···················· | ···················· | ····················· | ···················· | ···················· | ···················· | |
| D | 8727 hCoV-19/Canada/BC_25416870/2020\|EPI_ISL_463224\|202 | ···················· | ···················· | ····················· | ···················· | ···················· | ···················· | |
| D | 8728 hCoV-19/Canada/BC_25515215/2020\|EPI_ISL_463225\|202 | ···················· | ···················· | ····················· | ···················· | ···················· | ···················· | |
| D | 8729 hCoV-19/Canada/BC_25574199/2020\|EPI_ISL_463226\|202 | ···················· | ···················· | ····················· | ···················· | ···················· | ···················· | |
| D | 8730 hCoV-19/Canada/BC_25593681/2020\|EPI_ISL_463227\|202 | ···················· | ···················· | ····················· | ···················· | ···················· | ···················· | |
| D | 8731 hCoV-19/Canada/BC_25856970/2020\|EPI_ISL_463228\|202 | ···················· | ···················· | ····················· | ···················· | ···················· | ···················· | |
| D | 8732 hCoV-19/Canada/BC_25857092/2020\|EPI_ISL_463229\|202 | ···················· | ···················· | ····················· | ···················· | ···················· | ···················· | |
| D | 8733 hCoV-19/Canada/BC_26101196/2020\|EPI_ISL_463230\|202 | ···················· | ···················· | ····················· | ···················· | ···················· | ···················· | |
| D | 8734 hCoV-19/Canada/BC_26172976/2020\|EPI_ISL_463231\|202 | ···················· | ···················· | ····················· | ···················· | ···················· | ···················· | |
| D | 8735 hCoV-19/Canada/BC_26384651/2020\|EPI_ISL_463232\|202 | ···················· | ···················· | ····················· | ···················· | ···················· | ···················· | |
| D | 8736 hCoV-19/Canada/BC_26509737/2020\|EPI_ISL_463235\|202 | ···················· | ···················· | ····················· | ···················· | ···················· | ···················· | |
| D | 8737 hCoV-19/Canada/BC_26564890/2020\|EPI_ISL_463236\|202 | ···················· | ···················· | ····················· | ···················· | ···················· | ···················· | |
| D | 8738 hCoV-19/Canada/BC_26584470/2020\|EPI_ISL_463237\|202 | ···················· | ···················· | ····················· | ···················· | ···················· | ···················· | |
| D | 8739 hCoV-19/Canada/BC_26585549/2020\|EPI_ISL_463238\|202 | ···················· | ···················· | ····················· | ···················· | ···················· | ···················· | |
| D | 8740 hCoV-19/Canada/BC_26590014/2020\|EPI_ISL_463239\|202 | ···················· | ···················· | ····················· | ···················· | ···················· | ···················· | |
| D | 8741 hCoV-19/Canada/BC_26590284/2020\|EPI_ISL_463240\|202 | ···················· | ···················· | ····················· | ···················· | ···················· | ···················· | |
| D | 8742 hCoV-19/Canada/BC_26591461/2020\|EPI_ISL_463241\|202 | ···················· | ···················· | ····················· | ···················· | ···················· | ···················· | |
| D | 8743 hCoV-19/Canada/BC_26602426/2020\|EPI_ISL_463242\|202 | ···················· | ···················· | ····················· | ···················· | ···················· | ···················· | |
| D | 8744 hCoV-19/Canada/BC_26605771/2020\|EPI_ISL_463243\|202 | ···················· | ···················· | ····················· | ···················· | ···················· | ···················· | |
| D | 8745 hCoV-19/Canada/BC_26611727/2020\|EPI_ISL_463244\|202 | ···················· | ···················· | ····················· | ···················· | ···················· | ···················· | |
| D | 8748 hCoV-19/England/20109002804/2020\|EPI_ISL_464507\|20 | ···················· | ···················· | ····················· | ···················· | ···················· | ···················· | |
| D | 8749 hCoV-19/England/20109002904/2020\|EPI_ISL_464508\|20 | ···················· | ···················· | ····················· | ···················· | ···················· | ···················· | |
| D | 8751 hCoV-19/England/20109005704/2020\|EPI_ISL_464518\|20 | ···················· | ···················· | ····················· | ···················· | ···················· | ···················· | |
| D | 8752 hCoV-19/England/20109005904/2020\|EPI_ISL_464520\|20 | ···················· | ···················· | ····················· | ···················· | ···················· | ···················· | |
| D | 8753 hCoV-19/England/20109008404/2020\|EPI_ISL_464532\|20 | ···················· | ···················· | ····················· | ···················· | ···················· | ···················· | |
| D | 8754 hCoV-19/England/20109008604/2020\|EPI_ISL_464534\|20 | ···················· | ···················· | ····················· | ···················· | ···················· | ···················· | |
| D | 8755 hCoV-19/England/20109008704/2020\|EPI_ISL_464535\|20 | ···················· | ···················· | ····················· | ···················· | ···················· | ···················· | |
| D | 8756 hCoV-19/England/20109009804/2020\|EPI_ISL_464541\|20 | ···················· | ···················· | ····················· | ···················· | ···················· | ···················· | |
| D | 8757 hCoV-19/England/20109019704/2020\|EPI_ISL_464545\|20 | ···················· | ···················· | ····················· | ···················· | ···················· | ···················· | |
| D | 8758 hCoV-19/England/20109023404/2020\|EPI_ISL_464546\|20 | ···················· | ···················· | ····················· | ···················· | ···················· | ···················· | |
| D | 8759 hCoV-19/England/20109028804/2020\|EPI_ISL_464549\|20 | ···················· | ···················· | ····················· | ···················· | ···················· | ···················· | |
| D | 8760 hCoV-19/England/20109038504/2020\|EPI_ISL_464554\|20 | ···················· | ···················· | ····················· | ···················· | ···················· | ···················· | |
| D | 8764 hCoV-19/England/20109041904/2020\|EPI_ISL_464561\|20 | ···················· | ···················· | ····················· | ···················· | ···················· | ···················· | |
| D | 8765 hCoV-19/England/20109042004/2020\|EPI_ISL_464562\|20 | ···················· | ···················· | ····················· | ···················· | ···················· | ···················· | |

| | | | | | | | | |
|---|---|---|---|---|---|---|---|---|
| D | 8766 | hCoV-19/England/20109045404/2020\|EPI_ISL_464563\|20 | ···················· | ············A······· | ···················· | ···················· | ···················· | ···················· |
| D | 8767 | hCoV-19/England/20109047304/2020\|EPI_ISL_464567\|20 | ···················· | ···················· | ···················· | ···················· | ···················· | ···················· |
| D | 8768 | hCoV-19/England/20136031904/2020\|EPI_ISL_465820\|20 | ···················· | ···················· | ···················· | ···················· | ···················· | ···················· |
| D | 8769 | hCoV-19/England/20136102904/2020\|EPI_ISL_465821\|20 | ···················· | ···················· | ···················· | ···················· | ···················· | ···················· |
| D | 8770 | hCoV-19/England/20136103004/2020\|EPI_ISL_465822\|20 | ···················· | ···················· | ···················· | ···················· | ···················· | ···················· |
| D | 8771 | hCoV-19/England/20136103104/2020\|EPI_ISL_465823\|20 | ···················· | ···················· | ···················· | ···················· | ···················· | ···················· |
| D | 8772 | hCoV-19/England/20136103204/2020\|EPI_ISL_465824\|20 | ···················· | ···················· | ···················· | ···················· | ···················· | ···················· |
| D | 8773 | hCoV-19/England/201380235/2020\|EPI_ISL_465834\|2020 | ···················· | ···················· | ···················· | ···················· | ···················· | ···················· |
| D | 8774 | hCoV-19/England/20139071104/2020\|EPI_ISL_465864\|20 | ···················· | ···················· | ···················· | ···················· | ···················· | ···················· |
| D | 8775 | hCoV-19/England/20140001504/2020\|EPI_ISL_465865\|20 | ···················· | ···················· | ···················· | ···················· | ···················· | ···················· |
| D | 8776 | hCoV-19/England/20140002104/2020\|EPI_ISL_465866\|20 | ···················· | ···················· | ···················· | ···················· | ···················· | ···················· |
| D | 8777 | hCoV-19/England/20140002304/2020\|EPI_ISL_465867\|20 | ···················· | ···················· | ···················· | ···················· | ···················· | ···················· |
| D | 8778 | hCoV-19/England/20140002404/2020\|EPI_ISL_465868\|20 | ···················· | ···················· | ···················· | ···················· | ···················· | ···················· |
| D | 8779 | hCoV-19/England/20140002504/2020\|EPI_ISL_465869\|20 | ···················· | ···················· | ···················· | ···················· | ···················· | ···················· |
| D | 8780 | hCoV-19/England/20140003204/2020\|EPI_ISL_465870\|20 | ···················· | ···················· | ···················· | ···················· | ···················· | ···················· |
| D | 8781 | hCoV-19/England/20140004104/2020\|EPI_ISL_465871\|20 | ···················· | ···················· | ···················· | ···················· | ···················· | ···················· |
| D | 8782 | hCoV-19/England/20140004904/2020\|EPI_ISL_465872\|20 | ···················· | ···················· | ···················· | ···················· | ···················· | ···················· |
| D | 8783 | hCoV-19/England/20140005404/2020\|EPI_ISL_465873\|20 | ···················· | ···················· | ···················· | ···················· | ···················· | ···················· |
| D | 8784 | hCoV-19/England/20140006304/2020\|EPI_ISL_465874\|20 | ···················· | ···················· | ···················· | ···················· | ···················· | ···················· |
| D | 8785 | hCoV-19/England/20140006904/2020\|EPI_ISL_465875\|20 | ···················· | ···················· | ···················· | ···················· | ···················· | ···················· |
| D | 8786 | hCoV-19/England/20140007502/2020\|EPI_ISL_465876\|20 | ···················· | ···················· | ···················· | ···················· | ···················· | ···················· |
| D | 8787 | hCoV-19/England/20140008604/2020\|EPI_ISL_465877\|20 | ···················· | ···················· | ···················· | ···················· | ···················· | ···················· |
| D | 8788 | hCoV-19/England/20140008904/2020\|EPI_ISL_465878\|20 | ···················· | ···················· | ···················· | ···················· | ···················· | ···················· |
| D | 8789 | hCoV-19/England/20140009904/2020\|EPI_ISL_465879\|20 | ···················· | ···················· | ···················· | ···················· | ···················· | ···················· |
| D | 8790 | hCoV-19/England/20140060204/2020\|EPI_ISL_465881\|20 | ···················· | ···················· | ···················· | ···················· | ···················· | ···················· |
| D | 8791 | hCoV-19/England/20140063804/2020\|EPI_ISL_465883\|20 | ···················· | ···················· | ···················· | ···················· | ···················· | ···················· |
| D | 8792 | hCoV-19/England/20140064404/2020\|EPI_ISL_465884\|20 | ···················· | ···················· | ···················· | ···················· | ···················· | ···················· |
| D | 8793 | hCoV-19/England/20140064604/2020\|EPI_ISL_465885\|20 | ···················· | ···················· | ···················· | ···················· | ···················· | ···················· |
| D | 8794 | hCoV-19/England/20140068504/2020\|EPI_ISL_465887\|20 | ···················· | ···················· | ···················· | ···················· | ···················· | ···················· |
| D | 8795 | hCoV-19/England/20140068704/2020\|EPI_ISL_465888\|20 | ···················· | ···················· | ···················· | ···················· | ···················· | ···················· |
| D | 8796 | hCoV-19/England/20140073104/2020\|EPI_ISL_465890\|20 | ···················· | ···················· | ···················· | ···················· | ···················· | ···················· |
| D | 8797 | hCoV-19/England/20142001904/2020\|EPI_ISL_465895\|20 | ···················· | ···················· | ···················· | ···················· | ···················· | ···················· |
| D | 8798 | hCoV-19/England/20142003004/2020\|EPI_ISL_465896\|20 | ···················· | ···················· | ···················· | ···················· | ···················· | ···················· |
| D | 8799 | hCoV-19/Australia/SAP288/2020\|EPI_ISL_468020\|2020- | ···················· | ···················· | ···················· | ···················· | ···················· | ···················· |
| D | 8800 | hCoV-19/Australia/SAP289/2020\|EPI_ISL_468021\|2020- | ···················· | ············A······· | ···················· | ···················· | ···················· | ···················· |
| D | 8801 | hCoV-19/Australia/SAP263/2020\|EPI_ISL_468022\|2020- | ···················· | ···················· | ···················· | ···················· | ···················· | ···················· |
| D | 8802 | hCoV-19/Australia/SAP242/2020\|EPI_ISL_468023\|2020- | ···················· | ···················· | ···················· | ···················· | ···················· | ···················· |
| D | 8803 | hCoV-19/Australia/SAP226/2020\|EPI_ISL_468024\|2020- | ···················· | ···················· | ···················· | ···················· | ···················· | ···················· |
| D | 8804 | hCoV-19/Australia/SAP297/2020\|EPI_ISL_468025\|2020- | ···················· | ···················· | ···················· | ···················· | ···················· | ···················· |
| D | 8805 | hCoV-19/Australia/SAP221/2020\|EPI_ISL_468026\|2020- | ···················· | ···················· | ···················· | ···················· | ···················· | ···················· |
| D | 8806 | hCoV-19/Australia/SAP253/2020\|EPI_ISL_468027\|2020- | ···················· | ···················· | ···················· | ···················· | ···················· | ···················· |
| D | 8807 | hCoV-19/Australia/SAP256/2020\|EPI_ISL_468028\|2020- | ···················· | ···················· | ···················· | ···················· | ···················· | ···················· |
| D | 8808 | hCoV-19/Australia/SAP273/2020\|EPI_ISL_468029\|2020- | ···················· | ·A·················· | ···················· | ···················· | ···················· | ···················· |
| D | 8809 | hCoV-19/Australia/SAP294/2020\|EPI_ISL_468030\|2020- | ···················· | ···················· | ···················· | ···················· | ···················· | ···················· |
| D | 8810 | hCoV-19/Australia/SAP203/2020\|EPI_ISL_468031\|2020- | ···················· | ···················· | ···················· | ···················· | ···················· | ···················· |
| D | 8811 | hCoV-19/Australia/SAP214/2020\|EPI_ISL_468032\|2020- | ···················· | ···················· | ···················· | ···················· | ···················· | ···················· |
| D | 8812 | hCoV-19/Australia/SAP237/2020\|EPI_ISL_468033\|2020- | ···················· | ···················· | ···················· | ···················· | ···················· | ···················· |
| D | 8813 | hCoV-19/Australia/SAP293/2020\|EPI_ISL_468034\|2020- | ···················· | ···················· | ···················· | ···················· | ···················· | ···················· |
| D | 8814 | hCoV-19/Australia/SAP285/2020\|EPI_ISL_468035\|2020- | ···················· | ···················· | ···················· | ···················· | ···················· | ···················· |
| D | 8815 | hCoV-19/Australia/SAP206/2020\|EPI_ISL_468036\|2020- | ···················· | ···················· | ···················· | ···················· | ···················· | ···················· |
| D | 8816 | hCoV-19/Australia/SAP305/2020\|EPI_ISL_468037\|2020- | ···················· | ···················· | ···················· | ···················· | ···················· | ···················· |
| D | 8817 | hCoV-19/Australia/SAP099/2020\|EPI_ISL_468038\|2020- | ···················· | ···················· | ···················· | ···················· | ···················· | ···················· |
| D | 8818 | hCoV-19/Australia/SAP097/2020\|EPI_ISL_468039\|2020- | ···················· | ···················· | ···················· | ···················· | ···················· | ···················· |
| D | 8819 | hCoV-19/Australia/SAP271/2020\|EPI_ISL_468040\|2020- | ···················· | ···················· | ···················· | ···················· | ···················· | ···················· |
| D | 8820 | hCoV-19/Australia/SAP012/2020\|EPI_ISL_468041\|2020- | ···················· | ···················· | ···················· | ···················· | ···················· | ···················· |
| D | 8821 | hCoV-19/Australia/SAP270/2020\|EPI_ISL_468042\|2020- | ···················· | ···················· | ···················· | ···················· | ···················· | ···················· |
| D | 8822 | hCoV-19/Australia/SAP268/2020\|EPI_ISL_468043\|2020- | ···················· | ···················· | ···················· | ···················· | ···················· | ···················· |
| D | 8823 | hCoV-19/England/BRIS-12BBB2/2020\|EPI_ISL_469350\|20 | ···················· | ···················· | ···················· | ···················· | ···················· | ···················· |
| D | 8824 | hCoV-19/England/BRIS-12BC73/2020\|EPI_ISL_469353\|20 | ···················· | ···················· | ···················· | ···················· | ···················· | ···················· |
| D | 8825 | hCoV-19/England/BRIS-12BF01/2020\|EPI_ISL_469354\|20 | ···················· | ···················· | ···················· | ···················· | ···················· | ···················· |
| D | 8826 | hCoV-19/England/BRIS-12BC37/2020\|EPI_ISL_469355\|20 | ···················· | ···················· | ···················· | ···················· | ···················· | ···················· |
| D | 8827 | hCoV-19/England/BRIS-12BBA3/2020\|EPI_ISL_469356\|20 | ···················· | ···················· | ···················· | ···················· | ···················· | ···················· |
| D | 8828 | hCoV-19/England/BRIS-12B96D/2020\|EPI_ISL_469357\|20 | ···················· | ···················· | ···················· | ···················· | ···················· | ···················· |
| D | 8829 | hCoV-19/England/BRIS-12BCA0/2020\|EPI_ISL_469358\|20 | ···················· | ···················· | ···················· | ···················· | ···················· | ···················· |
| D | 8830 | hCoV-19/England/BRIS-12BA1F/2020\|EPI_ISL_469359\|20 | ···················· | ···················· | ···················· | ···················· | ···················· | ···················· |
| D | 8831 | hCoV-19/England/BRIS-12BFE3/2020\|EPI_ISL_469360\|20 | ···················· | ···················· | ···················· | ···················· | ···················· | ···················· |
| D | 8832 | hCoV-19/England/BRIS-12BF3E/2020\|EPI_ISL_469361\|20 | ···················· | ···················· | ···················· | ···················· | ···················· | ···················· |
| D | 8833 | hCoV-19/England/BRIS-12BB58/2020\|EPI_ISL_469362\|20 | ···················· | ···················· | ···················· | ···················· | ···················· | ···················· |
| D | 8834 | hCoV-19/England/BRIS-12BD8F/2020\|EPI_ISL_469364\|20 | ···················· | ···················· | ···················· | ···················· | ···················· | ···················· |
| D | 8835 | hCoV-19/England/BRIS-12C067/2020\|EPI_ISL_469365\|20 | ···················· | ···················· | ···················· | ···················· | ···················· | ···················· |
| D | 8836 | hCoV-19/England/BRIS-12C049/2020\|EPI_ISL_469366\|20 | ···················· | ···················· | ···················· | ···················· | ···················· | ···················· |
| D | 8837 | hCoV-19/England/BRIS-12BD70/2020\|EPI_ISL_469368\|20 | ···················· | ···················· | ···················· | ···················· | ···················· | ···················· |
| D | 8838 | hCoV-19/England/BRIS-12B8F7/2020\|EPI_ISL_469369\|20 | ···················· | ···A················ | ···················· | ···················· | ···················· | ···················· |
| D | 8839 | hCoV-19/England/BRIS-12C0B2/2020\|EPI_ISL_469370\|20 | ···················· | ···················· | ···················· | ···················· | ···················· | ···················· |
| D | 8840 | hCoV-19/England/BRIS-12BAA6/2020\|EPI_ISL_469372\|20 | ···················· | ···················· | ···················· | ···················· | ···················· | ···················· |
| D | 8841 | hCoV-19/England/BRIS-12C3B9/2020\|EPI_ISL_469373\|20 | ···················· | ···················· | ···················· | ···················· | ···················· | ···················· |
| D | 8842 | hCoV-19/England/BRIS-12BBD0/2020\|EPI_ISL_469375\|20 | ···················· | ···················· | ···················· | ···················· | ···················· | ···················· |
| D | 8843 | hCoV-19/England/BRIS-12C331/2020\|EPI_ISL_469376\|20 | ···················· | ···················· | ···················· | ···················· | ···················· | ···················· |

| | | | | | | | | |
|---|---|---|---|---|---|---|---|---|
| D | 8844 hCoV-19/England/BRIS-12C3C8/2020\|EPI_ISL_469377\|20 | .................... | .................... | .................... | .................... | .................... | .................... | |
| D | 8845 hCoV-19/England/BRIS-12BCBF/2020\|EPI_ISL_469379\|20 | .................... | .................... | .................... | .................... | .................... | .................... | |
| D | 8846 hCoV-19/England/BRIS-12C173/2020\|EPI_ISL_469380\|20 | .................... | .................... | .................... | .................... | .................... | .................... | |
| D | 8847 hCoV-19/England/BRIS-12C0FE/2020\|EPI_ISL_469381\|20 | .................... | .................... | .................... | .................... | .................... | .................... | |
| D | 8848 hCoV-19/England/BRIS-12BE04/2020\|EPI_ISL_469382\|20 | .................... | .................... | .................... | .................... | .................... | .................... | |
| D | 8849 hCoV-19/England/BRIS-12BF5C/2020\|EPI_ISL_469383\|20 | .................... | .................... | .................... | .................... | .................... | .................... | |
| D | 8850 hCoV-19/England/BRIS-12C0A3/2020\|EPI_ISL_469384\|20 | .................... | .................... | .................... | .................... | .................... | .................... | |
| D | 8851 hCoV-19/England/BRIS-12BE22/2020\|EPI_ISL_469385\|20 | .................... | .................... | .................... | .................... | .................... | .................... | |
| D | 8852 hCoV-19/England/BRIS-12BA97/2020\|EPI_ISL_469386\|20 | .................... | .................... | .................... | .................... | .................... | .................... | |
| D | 8853 hCoV-19/England/BRIS-12BB1C/2020\|EPI_ISL_469387\|20 | .................... | .................... | .................... | .................... | .................... | .................... | |
| D | 8854 hCoV-19/England/BRIS-12C340/2020\|EPI_ISL_469388\|20 | .................... | .................... | .................... | .................... | .................... | .................... | |
| D | 8855 hCoV-19/England/BRIS-12BBC1/2020\|EPI_ISL_469389\|20 | .................... | .................... | .................... | .................... | .................... | .................... | |
| D | 8856 hCoV-19/England/CAMB-8018D/2020\|EPI_ISL_470373\|202 | .................... | .................... | .................... | .................... | .................... | .................... | |
| D | 8857 hCoV-19/England/CAMB-7DD70/2020\|EPI_ISL_470374\|202 | .................... | .................... | .................... | .................... | .................... | .................... | |
| D | 8858 hCoV-19/England/CAMB-803A5/2020\|EPI_ISL_470375\|202 | .................... | .................... | .................... | .................... | .................... | .................... | |
| D | 8859 hCoV-19/England/CAMB-801E7/2020\|EPI_ISL_470379\|202 | .................... | .................... | .................... | .................... | .................... | .................... | |
| D | 8860 hCoV-19/England/CAMB-7DF10/2020\|EPI_ISL_470380\|202 | .................... | .................... | .................... | .................... | .................... | .................... | |
| D | 8861 hCoV-19/England/CAMB-7DDF8/2020\|EPI_ISL_470381\|202 | .................... | .................... | .................... | .................... | .................... | .................... | |
| D | 8862 hCoV-19/England/CAMB-80484/2020\|EPI_ISL_470382\|202 | .................... | .................... | .................... | .................... | .................... | .................... | |
| D | 8863 hCoV-19/England/CAMB-7DF3E/2020\|EPI_ISL_470383\|202 | .................... | .................... | .................... | .................... | .................... | .................... | |
| D | 8864 hCoV-19/England/CAMB-7BE51/2020\|EPI_ISL_470385\|202 | .................... | .................... | .................... | .................... | .................... | .................... | |
| D | 8865 hCoV-19/England/CAMB-7F6BF/2020\|EPI_ISL_470386\|202 | .................... | .................... | .................... | .................... | .................... | .................... | |
| D | 8866 hCoV-19/England/CAMB-800F9/2020\|EPI_ISL_470387\|202 | .................... | .................... | .................... | .................... | .................... | .................... | |
| D | 8867 hCoV-19/England/CAMB-80ACE/2020\|EPI_ISL_470388\|202 | .................... | .................... | .................... | .................... | .................... | .................... | |
| D | 8868 hCoV-19/England/CAMB-80624/2020\|EPI_ISL_470390\|202 | .................... | .................... | .................... | .................... | .................... | .................... | |
| D | 8869 hCoV-19/England/CAMB-800DB/2020\|EPI_ISL_470392\|202 | .................... | .................... | .................... | .................... | .................... | .................... | |
| D | 8870 hCoV-19/England/CAMB-7F7CB/2020\|EPI_ISL_470393\|202 | .................... | .................... | .................... | .................... | .................... | .................... | |
| D | 8871 hCoV-19/England/CAMB-7DDDA/2020\|EPI_ISL_470394\|202 | .................... | .................... | .................... | .................... | .................... | .................... | |
| D | 8872 hCoV-19/England/CAMB-7F5A3/2020\|EPI_ISL_470395\|202 | .................... | .................... | .................... | .................... | .................... | .................... | |
| D | 8873 hCoV-19/England/CAMB-7F50D/2020\|EPI_ISL_470397\|202 | .................... | .................... | .................... | .................... | .................... | .................... | |
| D | 8874 hCoV-19/England/CAMB-7F8B9/2020\|EPI_ISL_470398\|202 | .................... | .................... | .................... | .................... | .................... | .................... | |
| D | 8875 hCoV-19/USA/NE-11124/2020\|EPI_ISL_475200\|2020-05-0 | .................... | .................... | .................... | .................... | .................... | .................... | |
| D | 8876 hCoV-19/USA/NE-11163/2020\|EPI_ISL_475201\|2020-05-0 | .................... | .................... | .................... | .................... | .................... | .................... | |
| D | 8877 hCoV-19/USA/NE-11288/2020\|EPI_ISL_475203\|2020-05-0 | .................... | .................... | .................... | .................... | .................... | .................... | |
| D | 8878 hCoV-19/USA/NE-11317/2020\|EPI_ISL_475204\|2020-05-0 | .................... | .................... | .................... | .................... | .................... | .................... | |
| D | 8879 hCoV-19/USA/NE-11324/2020\|EPI_ISL_475205\|2020-05-0 | .................... | .................... | .................... | .................... | .................... | .................... | |
| D | 8880 hCoV-19/USA/NE-11331/2020\|EPI_ISL_475208\|2020-05-0 | .................... | .................... | .................... | .................... | .................... | .................... | |
| D | 8881 hCoV-19/USA/NE-11343/2020\|EPI_ISL_475209\|2020-05-0 | .................... | .................... | .................... | .................... | .................... | .................... | |
| D | 8882 hCoV-19/USA/NE-11435/2020\|EPI_ISL_475210\|2020-05-0 | .................... | .................... | .................... | .................... | .................... | .................... | |
| D | 8883 hCoV-19/USA/NE-11437/2020\|EPI_ISL_475211\|2020-05-0 | .................... | .................... | .................... | .................... | .................... | .................... | |
| D | 8884 hCoV-19/USA/NE-11445/2020\|EPI_ISL_475213\|2020-05-0 | .................... | .................... | .................... | .................... | .................... | .................... | |
| D | 8885 hCoV-19/USA/NE-11460/2020\|EPI_ISL_475214\|2020-05-0 | .................... | .................... | .................... | .................... | .................... | .................... | |
| D | 8886 hCoV-19/USA/NE-11462/2020\|EPI_ISL_475215\|2020-05-0 | .................... | .................... | .................... | .................... | .................... | .................... | |
| D | 8887 hCoV-19/USA/NE-11471/2020\|EPI_ISL_475216\|2020-05-0 | .................... | .................... | .................... | .................... | .................... | .................... | |
| D | 8888 hCoV-19/USA/NE-11480/2020\|EPI_ISL_475217\|2020-05-0 | .................... | .................... | .................... | .................... | .................... | .................... | |
| D | 8889 hCoV-19/USA/NE-11488/2020\|EPI_ISL_475219\|2020-05-0 | .................... | .................... | .................... | .................... | .................... | .................... | |
| D | 8890 hCoV-19/USA/NE-11503/2020\|EPI_ISL_475220\|2020-05-0 | .................... | .................... | .................... | .................... | .................... | .................... | |
| D | 8892 hCoV-19/USA/NE-11507/2020\|EPI_ISL_475222\|2020-05-0 | .................... | .................... | .................... | .................... | .................... | .................... | |
| D | 8893 hCoV-19/Canada/QC_AD6/2020\|EPI_ISL_476500\|2020-04- | .................... | .................... | .................... | .................... | .................... | .................... | |
| D | 8894 hCoV-19/Canada/QC_AD7/2020\|EPI_ISL_476501\|2020-04- | .................... | .................... | .................... | .................... | .................... | .................... | |
| D | 8895 hCoV-19/Canada/QC_AG3/2020\|EPI_ISL_476502\|2020-04- | .................... | .................... | .................... | .................... | .................... | .................... | |
| D | 8896 hCoV-19/Canada/QC_AG8/2020\|EPI_ISL_476503\|2020-04- | .................... | .................... | .................... | .................... | .................... | .................... | |
| D | 8897 hCoV-19/Canada/QC_AI6/2020\|EPI_ISL_476505\|2020-04- | .................... | .................... | .................... | .................... | .................... | .................... | |
| D | 8898 hCoV-19/Canada/QC_AJ8/2020\|EPI_ISL_476506\|2020-04- | .................... | .................... | .................... | .................... | .................... | .................... | |
| D | 8899 hCoV-19/Canada/QC_AK1/2020\|EPI_ISL_476507\|2020-04- | .................... | .................... | .................... | .................... | .................... | .................... | |
| D | 8900 hCoV-19/Canada/QC_AM1/2020\|EPI_ISL_476508\|2020-04- | .................... | .................... | .................... | .................... | .................... | .................... | |
| D | 8901 hCoV-19/Canada/QC_AM6/2020\|EPI_ISL_476509\|2020-04- | .................... | .................... | .................... | .................... | .................... | .................... | |
| D | 8902 hCoV-19/Canada/QC_AP8/2020\|EPI_ISL_476510\|2020-04- | .................... | .................... | .................... | .................... | .................... | .................... | |
| D | 8903 hCoV-19/USA/CT-Yale-293/2020\|EPI_ISL_476522\|2020-0 | .................... | .................... | .................... | .................... | .................... | .................... | |
| D | 8904 hCoV-19/USA/CT-Yale-294/2020\|EPI_ISL_476523\|2020-0 | .................... | .................... | .................... | .................... | .................... | .................... | |
| D | 8905 hCoV-19/USA/CT-Yale-296/2020\|EPI_ISL_476525\|2020-0 | .................... | .................... | .................... | .................... | .................... | .................... | |
| D | 8906 hCoV-19/USA/CT-Yale-302/2020\|EPI_ISL_476531\|2020-0 | .................... | .................... | .................... | .................... | .................... | .................... | |
| D | 8907 hCoV-19/USA/CT-Yale-314/2020\|EPI_ISL_476542\|2020-0 | .................... | .................... | .................... | .................... | .................... | .................... | |
| D | 8908 hCoV-19/USA/CT-Yale-317/2020\|EPI_ISL_476545\|2020-0 | .................... | .................... | .................... | .................... | .................... | .................... | |
| D | 8909 hCoV-19/USA/CT-Yale-319/2020\|EPI_ISL_476547\|2020-0 | .................... | .................... | .................... | .................... | .................... | .................... | |
| D | 8910 hCoV-19/USA/CT-Yale-320/2020\|EPI_ISL_476548\|2020-0 | .................... | ...................A | .................... | .................... | .................... | .................... | 1 |
| D | 8911 hCoV-19/USA/CT-Yale-321/2020\|EPI_ISL_476549\|2020-0 | .................... | .................... | .................... | .................... | .................... | .................... | |
| D | 8912 hCoV-19/USA/CT-Yale-322/2020\|EPI_ISL_476550\|2020-0 | .................... | .................... | .................... | .................... | .................... | .................... | |
| D | 8913 hCoV-19/USA/CT-Yale-324/2020\|EPI_ISL_476552\|2020-0 | .................... | .................... | .................... | .................... | .................... | .................... | |
| D | 8914 hCoV-19/USA/CT-Yale-325/2020\|EPI_ISL_476553\|2020-0 | .................... | .................... | .................... | .................... | .................... | .................... | |
| D | 8915 hCoV-19/USA/CT-Yale-326/2020\|EPI_ISL_476554\|2020-0 | .................... | .................... | .................... | .................... | .................... | .................... | |
| D | 8916 hCoV-19/England/CAMB-1B4F1C/2020\|EPI_ISL_477800\|20 | .................... | .................... | .................... | .................... | .................... | .................... | |
| D | 8917 hCoV-19/England/CAMB-1B4F49/2020\|EPI_ISL_477802\|20 | .................... | .................... | .................... | .................... | .................... | .................... | |
| D | 8918 hCoV-19/England/CAMB-1B4F94/2020\|EPI_ISL_477805\|20 | .................... | .................... | .................... | .................... | .................... | .................... | |
| D | 8919 hCoV-19/England/CAMB-1B4FA3/2020\|EPI_ISL_477806\|20 | .................... | .................... | .................... | .................... | .................... | .................... | |
| D | 8920 hCoV-19/England/CAMB-1B4FB2/2020\|EPI_ISL_477807\|20 | .................... | .................... | .................... | .................... | .................... | .................... | |
| D | 8921 hCoV-19/England/CAMB-1B4FC1/2020\|EPI_ISL_477808\|20 | .................... | .................... | .................... | .................... | .................... | .................... | |
| D | 8922 hCoV-19/England/CAMB-1B4FD0/2020\|EPI_ISL_477809\|20 | .................... | .................... | .................... | .................... | .................... | .................... | |

| | | | | | | | | | |
|---|---|---|---|---|---|---|---|---|---|
| D | 8923 | hCoV-19/England/CAMB-1B4FFE/2020\|EPI_ISL_477810\|20 | .................... | .................... | .................... | .................... | .................... | .................... | |
| D | 8924 | hCoV-19/England/CAMB-1B5018/2020\|EPI_ISL_477811\|20 | .................... | .................... | .................... | .................... | .................... | .................... | |
| D | 8925 | hCoV-19/England/CAMB-1B5027/2020\|EPI_ISL_477812\|20 | .................... | .................... | .................... | .................... | .................... | .................... | |
| D | 8926 | hCoV-19/England/CAMB-1B5036/2020\|EPI_ISL_477813\|20 | .................... | .................... | .................... | .................... | .................... | .................... | |
| D | 8927 | hCoV-19/England/CAMB-1B50BE/2020\|EPI_ISL_477814\|20 | .................... | .................... | .................... | .................... | .................... | .................... | |
| D | 8928 | hCoV-19/Scotland/CVR1509/2020\|EPI_ISL_477822\|2020- | .................... | ...................A | .................... | .................... | .................... | .................... | 1 |
| D | 8929 | hCoV-19/Scotland/CVR1517/2020\|EPI_ISL_477823\|2020- | .................... | .................... | .................... | .................... | .................... | .................... | |
| D | 8930 | hCoV-19/Scotland/CVR1883/2020\|EPI_ISL_477825\|2020- | .................... | .................... | .................... | .................... | .................... | .................... | |
| D | 8931 | hCoV-19/Scotland/CVR1897/2020\|EPI_ISL_477827\|2020- | .................... | ...................A | .................... | .................... | .................... | .................... | 1 |
| D | 8932 | hCoV-19/Scotland/CVR1898/2020\|EPI_ISL_477828\|2020- | .................... | .................... | .................... | .................... | .................... | .................... | |
| D | 8933 | hCoV-19/Scotland/CVR1900/2020\|EPI_ISL_477829\|2020- | .................... | .................... | .................... | .................... | .................... | .................... | |
| D | 8934 | hCoV-19/Scotland/CVR1901/2020\|EPI_ISL_477830\|2020- | .................... | .................... | .................... | .................... | .................... | .................... | |
| D | 8935 | hCoV-19/Scotland/CVR1903/2020\|EPI_ISL_477831\|2020- | .................... | .................... | .................... | .................... | .................... | .................... | |
| D | 8936 | hCoV-19/Scotland/CVR1905/2020\|EPI_ISL_477833\|2020- | .................... | ...................A | .................... | .................... | .................... | .................... | 1 |
| D | 8937 | hCoV-19/Scotland/CVR1911/2020\|EPI_ISL_477834\|2020- | .................... | .................... | .................... | .................... | .................... | .................... | |
| D | 8938 | hCoV-19/Scotland/CVR1940/2020\|EPI_ISL_477838\|2020- | .................... | .................... | .................... | .................... | .................... | .................... | |
| D | 8939 | hCoV-19/Scotland/CVR1963/2020\|EPI_ISL_477839\|2020- | .................... | .............A...... | .................... | .................... | .................... | .................... | |
| D | 8940 | hCoV-19/Scotland/CVR1981/2020\|EPI_ISL_477841\|2020- | .................... | .................... | .................... | .................... | .................... | .................... | |
| D | 8941 | hCoV-19/Scotland/CVR2089/2020\|EPI_ISL_477853\|2020- | .................... | .................... | .................... | .................... | .................... | .................... | |
| D | 8942 | hCoV-19/Scotland/CVR2093/2020\|EPI_ISL_477855\|2020- | .................... | ...................A | .................... | .................... | .................... | .................... | 1 |
| D | 8943 | hCoV-19/Scotland/CVR2117/2020\|EPI_ISL_477856\|2020- | ..T................. | ...................A | .................... | .................... | .................... | .................... | 1 |
| D | 8944 | hCoV-19/Scotland/CVR2138/2020\|EPI_ISL_477861\|2020- | .................... | .................... | .................... | .................... | .................... | .................... | |
| D | 8945 | hCoV-19/Scotland/CVR2140/2020\|EPI_ISL_477862\|2020- | .................... | .................... | .................... | .................... | .................... | .................... | |
| D | 8946 | hCoV-19/Scotland/CVR2166/2020\|EPI_ISL_477866\|2020- | .................... | .................... | .................... | .................... | .................... | .................... | |
| D | 8947 | hCoV-19/Scotland/CVR2176/2020\|EPI_ISL_477867\|2020- | .................... | ...................A | .................... | .................... | .................... | .................... | 1 |
| D | 8948 | hCoV-19/Scotland/CVR2185/2020\|EPI_ISL_477869\|2020- | .................... | .................... | .................... | .................... | .................... | .................... | |
| D | 8949 | hCoV-19/Scotland/CVR2187/2020\|EPI_ISL_477870\|2020- | .................... | .................... | .................... | .................... | .................... | .................... | |
| D | 8950 | hCoV-19/Scotland/CVR2190/2020\|EPI_ISL_477871\|2020- | .................... | .................... | .................... | .................... | .................... | .................... | |
| D | 8951 | hCoV-19/Scotland/CVR2191/2020\|EPI_ISL_477872\|2020- | .................... | ...................A | .................... | .................... | .................... | .................... | 1 |
| D | 8952 | hCoV-19/Scotland/CVR2213/2020\|EPI_ISL_477874\|2020- | .................... | .................... | .................... | .................... | .................... | .................... | |
| D | 8953 | hCoV-19/Scotland/CVR2220/2020\|EPI_ISL_477875\|2020- | .................... | .................... | .................... | .................... | .................... | .................... | |
| D | 8954 | hCoV-19/Scotland/CVR2231/2020\|EPI_ISL_477876\|2020- | .................... | .................... | .................... | .................... | .................... | .................... | |
| D | 8955 | hCoV-19/Scotland/CVR2234/2020\|EPI_ISL_477877\|2020- | .................... | .A.................. | .................... | .................... | .................... | .................... | |
| D | 8956 | hCoV-19/Scotland/CVR2238/2020\|EPI_ISL_477879\|2020- | .................... | .................... | .................... | .................... | .................... | .................... | |
| D | 8957 | hCoV-19/Scotland/CVR2240/2020\|EPI_ISL_477881\|2020- | .................... | .................... | .................... | .................... | .................... | .................... | |
| D | 8958 | hCoV-19/Scotland/CVR2247/2020\|EPI_ISL_477883\|2020- | .................... | .................... | .................... | .................... | .................... | .................... | |
| D | 8959 | hCoV-19/Scotland/CVR2250/2020\|EPI_ISL_477884\|2020- | .................... | ...................A | .................... | .................... | .................... | .................... | 1 |
| D | 8960 | hCoV-19/Scotland/CVR2251/2020\|EPI_ISL_477885\|2020- | .................... | .................... | .................... | .................... | .................... | .................... | |
| D | 8961 | hCoV-19/USA/WA-S898/2020\|EPI_ISL_463388\|2020-04-27 | .................... | .................... | .................... | .................... | .................... | .................... | |
| D | 8962 | hCoV-19/USA/WA-S899/2020\|EPI_ISL_463389\|2020-04-28 | .................... | .................... | .................... | .................... | .................... | .................... | |
| D | 8963 | hCoV-19/USA/WA-S900/2020\|EPI_ISL_463390\|2020-04-28 | .................... | .................... | .................... | .................... | .................... | .................... | |
| D | 8964 | hCoV-19/USA/WA-S901/2020\|EPI_ISL_463391\|2020-04-28 | .................... | .................... | .................... | .................... | .................... | .................... | |
| D | 8965 | hCoV-19/USA/WA-S902/2020\|EPI_ISL_463392\|2020-04-22 | .................... | .................... | .................... | .................... | .................... | .................... | |
| D | 8966 | hCoV-19/USA/WA-S904/2020\|EPI_ISL_463394\|2020-04-27 | .................... | .................... | .................... | .................... | .................... | .................... | |
| D | 8967 | hCoV-19/USA/WA-S905/2020\|EPI_ISL_463395\|2020-04-27 | .................... | .................... | .................... | .................... | .................... | .................... | |
| D | 8968 | hCoV-19/USA/WA-S906/2020\|EPI_ISL_463396\|2020-04-27 | .................... | .................... | .................... | .................... | .................... | .................... | |
| D | 8969 | hCoV-19/USA/WA-S907/2020\|EPI_ISL_463397\|2020-04-27 | .................... | .................... | .................... | .................... | .................... | .................... | |
| D | 8970 | hCoV-19/USA/WA-S908/2020\|EPI_ISL_463398\|2020-04-27 | .................... | .................... | .................... | .................... | .................... | .................... | |
| D | 8971 | hCoV-19/USA/WA-S909/2020\|EPI_ISL_463399\|2020-04-27 | .................... | .................... | .................... | .................... | .................... | .................... | |
| D | 8972 | hCoV-19/England/20129067504/2020\|EPI_ISL_465808\|20 | .................... | .................... | .................... | .................... | .................... | .................... | |
| D | 8975 | hCoV-19/England/20136016104/2020\|EPI_ISL_465818\|20 | .................... | .................... | .................... | .................... | .................... | .................... | |
| D | 8976 | hCoV-19/England/20136016204/2020\|EPI_ISL_465819\|20 | .................... | .................... | .................... | .................... | .................... | .................... | |
| D | 8977 | hCoV-19/Australia/SAP216/2020\|EPI_ISL_468000\|2020- | .................... | .................... | .................... | .................... | .................... | .................... | |
| D | 8978 | hCoV-19/Australia/SAP215/2020\|EPI_ISL_468001\|2020- | .................... | .................... | .................... | .................... | .................... | .................... | |
| D | 8979 | hCoV-19/Australia/SAP244/2020\|EPI_ISL_468002\|2020- | .................... | .................... | .................... | .................... | .................... | .................... | |
| D | 8980 | hCoV-19/Australia/SAP207/2020\|EPI_ISL_468003\|2020- | .................... | .................... | .................... | .................... | .................... | .................... | |
| D | 8981 | hCoV-19/Australia/SAP284/2020\|EPI_ISL_468004\|2020- | .................... | .................... | .................... | .................... | .................... | .................... | |
| D | 8982 | hCoV-19/Australia/SAP236/2020\|EPI_ISL_468005\|2020- | .................... | .................... | .................... | .................... | .................... | .................... | |
| D | 8983 | hCoV-19/Australia/SAP302/2020\|EPI_ISL_468006\|2020- | .................... | .................... | .................... | .................... | .................... | .................... | |
| D | 8984 | hCoV-19/Australia/SAP298/2020\|EPI_ISL_468007\|2020- | .................... | .................... | .................... | .................... | .................... | .................... | |
| D | 8985 | hCoV-19/Australia/SAP229/2020\|EPI_ISL_468008\|2020- | .................... | .................... | .................... | .................... | .................... | .................... | |
| D | 8986 | hCoV-19/Australia/SAP027/2020\|EPI_ISL_468009\|2020- | .................... | .................... | .................... | .................... | .................... | .................... | |
| D | 8987 | hCoV-19/Australia/SAP257/2020\|EPI_ISL_468010\|2020- | .................... | .................... | .................... | .................... | .................... | .................... | |
| D | 8988 | hCoV-19/Australia/SAP022/2020\|EPI_ISL_468011\|2020- | .................... | .................... | .................... | .................... | .................... | .................... | |
| D | 8989 | hCoV-19/Australia/SAP017/2020\|EPI_ISL_468012\|2020- | .................... | .................... | .................... | .................... | .................... | .................... | |
| D | 8990 | hCoV-19/Australia/SAP204/2020\|EPI_ISL_468013\|2020- | .................... | .................... | .................... | .................... | .................... | .................... | |
| D | 8991 | hCoV-19/Australia/SAP246/2020\|EPI_ISL_468014\|2020- | .................... | .................... | .................... | .................... | .................... | .................... | |
| D | 8992 | hCoV-19/Australia/SAP243/2020\|EPI_ISL_468015\|2020- | .................... | .................... | .................... | .................... | .................... | .................... | |
| D | 8993 | hCoV-19/Australia/SAP112/2020\|EPI_ISL_468016\|2020- | .................... | .................... | .................... | .................... | .................... | .................... | |
| D | 8994 | hCoV-19/Australia/SAP019/2020\|EPI_ISL_468017\|2020- | .................... | .................... | .................... | .................... | .................... | .................... | |
| D | 8995 | hCoV-19/Australia/SAP269/2020\|EPI_ISL_468018\|2020- | .................... | .................... | .................... | .................... | .................... | .................... | |
| D | 8996 | hCoV-19/Australia/SAP014/2020\|EPI_ISL_468019\|2020- | .................... | .................... | .................... | .................... | .................... | .................... | |
| D | 8997 | hCoV-19/Turkey/IMU-SP-03/2020\|EPI_ISL_468066\|2020- | .................... | .................... | .................... | .................... | .................... | .................... | |
| D | 8998 | hCoV-19/Bangladesh/CHRF-0002/2020\|EPI_ISL_468070\|2 | .................... | .................... | .................... | .................... | .................... | .................... | |
| D | 8999 | hCoV-19/Bangladesh/CHRF-0003/2020\|EPI_ISL_468071\|2 | .................... | .................... | .................... | .................... | .................... | .................... | |
| D | 9000 | hCoV-19/Bangladesh/CHRF-0004/2020\|EPI_ISL_468072\|2 | .................... | .................... | .................... | .................... | .................... | .................... | |
| D | 9001 | hCoV-19/Bangladesh/CHRF-0005/2020\|EPI_ISL_468073\|2 | .................... | .................... | .................... | .................... | .................... | .................... | |
| D | 9002 | hCoV-19/Bangladesh/CHRF-0007/2020\|EPI_ISL_468075\|2 | .................... | .................... | .................... | .................... | .................... | .................... | |

| | | | | | | | | |
|---|---|---|---|---|---|---|---|---|
| D | 9003 | hCoV-19/Bangladesh/CHRF-0008/2020\|EPI_ISL_468076\|2 | ···················· | ···················· | ···················· | ···················· | ···················· | ···················· |
| D | 9004 | hCoV-19/Bangladesh/CHRF-0009/2020\|EPI_ISL_468077\|2 | ···················· | ···················· | ···················· | ···················· | ···················· | ···················· |
| D | 9005 | hCoV-19/Bangladesh/CHRF-0010/2020\|EPI_ISL_468078\|2 | ···················· | ···················· | ···················· | ···················· | ···················· | ···················· |
| D | 9006 | hCoV-19/USA/WA-OHSU-0032/2020\|EPI_ISL_468081\|2020- | ···················· | ···················· | ···················· | ···················· | ···················· | ···················· |
| D | 9007 | hCoV-19/USA/WA-OHSU-0034/2020\|EPI_ISL_468082\|2020- | ···················· | ···················· | ···················· | ···················· | ···················· | ···················· |
| D | 9008 | hCoV-19/USA/WA-OHSU-0035/2020\|EPI_ISL_468083\|2020- | ···················· | ···················· | ···················· | ···················· | ···················· | ···················· |
| D | 9009 | hCoV-19/USA/WA-OHSU-0036/2020\|EPI_ISL_468084\|2020- | ···················· | ···················· | ···················· | ···················· | ···················· | ···················· |
| D | 9010 | hCoV-19/USA/OR-OHSU-0058/2020\|EPI_ISL_468089\|2020- | ···················· | ···················· | ···················· | ···················· | ···················· | ···················· |
| D | 9011 | hCoV-19/USA/OR-OHSU-0075/2020\|EPI_ISL_468095\|2020- | ···················· | ···················· | ···················· | ···················· | ···················· | ···················· |
| D | 9012 | hCoV-19/Bangladesh/BCSIR-NILMRC-091/2020\|EPI_ISL_4 | ···················· | ···················· | ···················· | ···················· | ···················· | ···················· |
| D | 9013 | hCoV-19/England/NORT-28B1F9/2020\|EPI_ISL_469302\|20 | ···················· | ···················· | ···················· | ···················· | ···················· | ···················· |
| D | 9014 | hCoV-19/England/NORT-29878E/2020\|EPI_ISL_469303\|20 | ···················· | ···················· | ···················· | ···················· | ···················· | ···················· |
| D | 9015 | hCoV-19/England/NORT-28B171/2020\|EPI_ISL_469307\|20 | ···················· | ···················· | ···················· | ···················· | ···················· | ···················· |
| D | 9016 | hCoV-19/England/NORT-28B223/2020\|EPI_ISL_469309\|20 | ···················· | ···················· | ···················· | ···················· | ···················· | ···················· |
| D | 9017 | hCoV-19/England/NORT-29879D/2020\|EPI_ISL_469312\|20 | ···················· | ···················· | ···················· | ···················· | ···················· | ···················· |
| D | 9018 | hCoV-19/England/NORT-28C87B/2020\|EPI_ISL_469313\|20 | ···················· | ···················· | ···················· | ···················· | ···················· | ···················· |
| D | 9019 | hCoV-19/England/NORT-28B153/2020\|EPI_ISL_469314\|20 | ···················· | ···················· | ···················· | ···················· | ···················· | ···················· |
| D | 9020 | hCoV-19/England/NORT-28C714/2020\|EPI_ISL_469315\|20 | ···················· | ···················· | ···················· | ···················· | ···················· | ···················· |
| D | 9021 | hCoV-19/England/NORT-298751/2020\|EPI_ISL_469316\|20 | ···················· | ···················· | ···················· | ···················· | ···················· | ···················· |
| D | 9022 | hCoV-19/England/NORT-28C86C/2020\|EPI_ISL_469319\|20 | ···················· | ···················· | ···················· | ···················· | ···················· | ···················· |
| D | 9023 | hCoV-19/England/NORT-28C9A5/2020\|EPI_ISL_469324\|20 | ···················· | ···················· | ···················· | ···················· | ···················· | ···················· |
| D | 9024 | hCoV-19/England/NORT-28C7AB/2020\|EPI_ISL_469325\|20 | ···················· | ···················· | ···················· | ···················· | ···················· | ···················· |
| D | 9025 | hCoV-19/England/NORT-28CA2A/2020\|EPI_ISL_469328\|20 | ···················· | ···················· | ···················· | ···················· | ···················· | ···················· |
| D | 9026 | hCoV-19/England/NORT-28CB81/2020\|EPI_ISL_469329\|20 | ···················· | ···················· | ···················· | ···················· | ···················· | ···················· |
| D | 9027 | hCoV-19/England/NORT-28CAFD/2020\|EPI_ISL_469330\|20 | ···················· | ···················· | ···················· | ···················· | ···················· | ···················· |
| D | 9028 | hCoV-19/England/NORT-28C69F/2020\|EPI_ISL_469335\|20 | ···················· | ···················· | ···················· | ···················· | ···················· | ···················· |
| D | 9029 | hCoV-19/England/NORT-28C6BD/2020\|EPI_ISL_469336\|20 | ···················· | ···················· | ···················· | ···················· | ···················· | ···················· |
| D | 9030 | hCoV-19/England/NORT-28C90F/2020\|EPI_ISL_469340\|20 | ···················· | ···················· | ···················· | ···················· | ···················· | ···················· |
| D | 9031 | hCoV-19/England/NORT-28C5DE/2020\|EPI_ISL_469344\|20 | ···················· | ···················· | ···················· | ···················· | ···················· | ···················· |
| D | 9032 | hCoV-19/England/BRIS-12BFD4/2020\|EPI_ISL_469345\|20 | ···················· | ···················· | ···················· | ···················· | ···················· | ···················· |
| D | 9033 | hCoV-19/England/BRIS-12BE8C/2020\|EPI_ISL_469346\|20 | ···················· | ···················· | ···················· | ···················· | ···················· | ···················· |
| D | 9034 | hCoV-19/England/BRIS-12B8D9/2020\|EPI_ISL_469347\|20 | ···················· | ···················· | ···················· | ···················· | ···················· | ···················· |
| D | 9035 | hCoV-19/England/BRIS-12C094/2020\|EPI_ISL_469348\|20 | ···················· | ···················· | ···················· | ···················· | ···················· | ···················· |
| D | 9036 | hCoV-19/England/BRIS-12C10A/2020\|EPI_ISL_469349\|20 | ···················· | ···················· | ···················· | ···················· | ···················· | ···················· |
| D | 9037 | hCoV-19/England/BRIS-12BCDD/2020\|EPI_ISL_469390\|20 | ···················· | ···················· | ···················· | ···················· | ···················· | ···················· |
| D | 9038 | hCoV-19/England/BRIS-12BD07/2020\|EPI_ISL_469391\|20 | ···················· | ···················· | ···················· | ···················· | ···················· | ···················· |
| D | 9039 | hCoV-19/England/BRIS-12BA4C/2020\|EPI_ISL_469392\|20 | ···················· | ···················· | ···················· | ···················· | ···················· | ···················· |
| D | 9040 | hCoV-19/England/BRIS-12B9F4/2020\|EPI_ISL_469394\|20 | ···················· | ···················· | ···················· | ···················· | ···················· | ···················· |
| D | 9041 | hCoV-19/England/BRIS-12BA2E/2020\|EPI_ISL_469395\|20 | ···················· | ···················· | ···················· | ···················· | ···················· | ···················· |
| D | 9042 | hCoV-19/England/BRIS-12C1FB/2020\|EPI_ISL_469396\|20 | ···················· | ···················· | ···················· | ···················· | ···················· | ···················· |
| D | 9043 | hCoV-19/England/BRIS-12BD43/2020\|EPI_ISL_469397\|20 | ···················· | ···················· | ···················· | ···················· | ···················· | ···················· |
| D | 9044 | hCoV-19/England/BRIS-12BA6A/2020\|EPI_ISL_469398\|20 | ···················· | ···················· | ···················· | ···················· | ···················· | ···················· |
| D | 9045 | hCoV-19/England/CAMB-1ABBD8/2020\|EPI_ISL_470300\|20 | ···················· | ···················· | ···················· | ···················· | ···················· | ···················· |
| D | 9046 | hCoV-19/England/CAMB-1AB26A/2020\|EPI_ISL_470302\|20 | ···················· | ···················· | ···················· | ···················· | ···················· | ···················· |
| D | 9047 | hCoV-19/England/CAMB-1AA6BA/2020\|EPI_ISL_470303\|20 | ···················· | ···················· | ···················· | ···················· | ···················· | ···················· |
| D | 9048 | hCoV-19/England/CAMB-1AABCA/2020\|EPI_ISL_470307\|20 | ···················· | ···················· | ···················· | ···················· | ···················· | ···················· |
| D | 9049 | hCoV-19/England/CAMB-1ABACC/2020\|EPI_ISL_470309\|20 | ···················· | ···················· | ···················· | ···················· | ···················· | ···················· |
| D | 9050 | hCoV-19/England/CAMB-1AAA18/2020\|EPI_ISL_470310\|20 | ···················· | ···················· | ···················· | ···················· | ···················· | ···················· |
| D | 9051 | hCoV-19/England/CAMB-1AA720/2020\|EPI_ISL_470311\|20 | ···················· | ···················· | ···················· | ···················· | ······T············· | ···················· |
| D | 9052 | hCoV-19/England/CAMB-1AB482/2020\|EPI_ISL_470312\|20 | ···················· | ···················· | ···················· | ···················· | ···················· | ···················· |
| D | 9053 | hCoV-19/England/CAMB-1AB1B8/2020\|EPI_ISL_470313\|20 | ···················· | ···················· | ···················· | ···················· | ···················· | ···················· |
| D | 9054 | hCoV-19/England/CAMB-1AA8C3/2020\|EPI_ISL_470314\|20 | ···················· | ···················· | ···················· | ···················· | ···················· | ···················· |
| D | 9055 | hCoV-19/England/CAMB-1ABDD2/2020\|EPI_ISL_470315\|20 | ···················· | ···················· | ···················· | ···················· | ···················· | ···················· |
| D | 9056 | hCoV-19/England/CAMB-1AB3FE/2020\|EPI_ISL_470316\|20 | ···················· | ···················· | ···················· | ···················· | ···················· | ···················· |
| D | 9057 | hCoV-19/England/CAMB-1AB9DE/2020\|EPI_ISL_470319\|20 | ···················· | ···················· | ···················· | ···················· | ···················· | ···················· |
| D | 9058 | hCoV-19/England/CAMB-1ABD5A/2020\|EPI_ISL_470320\|20 | ···················· | ···················· | ···················· | ···················· | ···················· | ···················· |
| D | 9059 | hCoV-19/England/CAMB-1ABC5D/2020\|EPI_ISL_470323\|20 | ···················· | ···················· | ···················· | ···················· | ···················· | ···················· |
| D | 9060 | hCoV-19/England/CAMB-1AB6B9/2020\|EPI_ISL_470324\|20 | ···················· | ···················· | ···················· | ···················· | ···················· | ···················· |
| D | 9061 | hCoV-19/England/CAMB-1ABC4E/2020\|EPI_ISL_470325\|20 | ···················· | ···················· | ···················· | ···················· | ···················· | ···················· |
| D | 9062 | hCoV-19/England/CAMB-1ABA44/2020\|EPI_ISL_470328\|20 | ···················· | ···················· | ···················· | ···················· | ···················· | ···················· |
| D | 9063 | hCoV-19/England/CAMB-1AB4BF/2020\|EPI_ISL_470329\|20 | ···················· | ···················· | ···················· | ···················· | ···················· | ···················· |
| D | 9064 | hCoV-19/England/CAMB-1AACA9/2020\|EPI_ISL_470330\|20 | ···················· | ···················· | ···················· | ···················· | ···················· | ···················· |
| D | 9065 | hCoV-19/England/CAMB-1AAB7F/2020\|EPI_ISL_470331\|20 | ···················· | ···················· | ···················· | ···················· | ···················· | ···················· |
| D | 9066 | hCoV-19/England/CAMB-1AB65F/2020\|EPI_ISL_470332\|20 | ···················· | ···················· | ···················· | ···················· | ···················· | ···················· |
| D | 9067 | hCoV-19/England/CAMB-1AB19A/2020\|EPI_ISL_470334\|20 | ···················· | ···················· | ···················· | ···················· | ···················· | ···················· |
| D | 9068 | hCoV-19/England/CAMB-1ABD69/2020\|EPI_ISL_470335\|20 | ···················· | ···················· | ···················· | ···················· | ···················· | ···················· |
| D | 9069 | hCoV-19/England/CAMB-1AA81E/2020\|EPI_ISL_470336\|20 | ···················· | ···················· | ···················· | ···················· | ···················· | ···················· |
| D | 9070 | hCoV-19/England/CAMB-1AAA54/2020\|EPI_ISL_470340\|20 | ···················· | ···················· | ···················· | ···················· | ···················· | ···················· |
| D | 9071 | hCoV-19/England/CAMB-1ABA80/2020\|EPI_ISL_470341\|20 | ···················· | ···················· | ···················· | ···················· | ···················· | ···················· |
| D | 9072 | hCoV-19/England/CAMB-1AB69B/2020\|EPI_ISL_470342\|20 | ···················· | ···················· | ···················· | ···················· | ···················· | ···················· |
| D | 9073 | hCoV-19/England/CAMB-1AB464/2020\|EPI_ISL_470343\|20 | ···················· | ···················· | ···················· | ···················· | ···················· | ···················· |
| D | 9074 | hCoV-19/England/CAMB-1AADB5/2020\|EPI_ISL_470344\|20 | ···················· | ···················· | ···················· | ···················· | ···················· | ···················· |
| D | 9075 | hCoV-19/England/CAMB-1ABC20/2020\|EPI_ISL_470345\|20 | ···················· | ···················· | ···················· | ···················· | ···················· | ···················· |
| D | 9076 | hCoV-19/England/CAMB-1ABC8A/2020\|EPI_ISL_470347\|20 | ···················· | ···················· | ···················· | ···················· | ···················· | ···················· |
| D | 9077 | hCoV-19/England/CAMB-1AB570/2020\|EPI_ISL_470348\|20 | ···················· | ···················· | ···················· | ···················· | ···················· | ···················· |
| D | 9078 | hCoV-19/England/CAMB-1AB6AA/2020\|EPI_ISL_470349\|20 | ···················· | ···················· | ···················· | ···················· | ···················· | ···················· |
| D | 9079 | hCoV-19/England/CAMB-1AAA27/2020\|EPI_ISL_470350\|20 | ···················· | ···················· | ···················· | ···················· | ···················· | ···················· |
| D | 9080 | hCoV-19/England/CAMB-1AAA72/2020\|EPI_ISL_470351\|20 | ···················· | ·A·················· | ···················· | ···················· | ···················· | ···················· |

| | | | | | | | |
|---|---|---|---|---|---|---|---|
| D | 9081 hCoV-19/England/CAMB-835F7/2020\|EPI_ISL_470353\|202 | ···················· | ···················· | ···················· | ···················· | ···················· | ···················· |
| D | 9082 hCoV-19/England/BRIS-124CC5/2020\|EPI_ISL_470356\|20 | ···················· | ···················· | ···················· | ···················· | ···················· | ···················· |
| D | 9083 hCoV-19/England/BRIS-124885/2020\|EPI_ISL_470357\|20 | ···················· | ···················· | ···················· | ···················· | ···················· | ···················· |
| D | 9084 hCoV-19/England/CAMB-7F44C/2020\|EPI_ISL_470359\|202 | ···················· | ···················· | ···················· | ···················· | ···················· | ···················· |
| D | 9085 hCoV-19/England/CAMB-7EAB4/2020\|EPI_ISL_470360\|202 | ···················· | ···················· | ···················· | ···················· | ···················· | ···················· |
| D | 9086 hCoV-19/England/CAMB-8078B/2020\|EPI_ISL_470361\|202 | ···················· | ···················· | ···················· | ···················· | ···················· | ···················· |
| D | 9087 hCoV-19/England/CAMB-7FA86/2020\|EPI_ISL_470363\|202 | ···················· | ···················· | ···················· | ···················· | ···················· | ···················· |
| D | 9088 hCoV-19/England/CAMB-8019C/2020\|EPI_ISL_470364\|202 | ···················· | ···················· | ···················· | ···················· | ···················· | ···················· |
| D | 9089 hCoV-19/England/CAMB-80448/2020\|EPI_ISL_470366\|202 | ···················· | ···················· | ···················· | ···················· | ···················· | ···················· |
| D | 9090 hCoV-19/England/CAMB-807D6/2020\|EPI_ISL_470367\|202 | ···················· | ···················· | ···················· | ···················· | ···················· | ···················· |
| D | 9091 hCoV-19/England/CAMB-7DE5F/2020\|EPI_ISL_470368\|202 | ···················· | ···················· | ···················· | ···················· | ···················· | ···················· |
| D | 9092 hCoV-19/England/CAMB-80660/2020\|EPI_ISL_470370\|202 | ···················· | ···················· | ···················· | ···················· | ···················· | ···················· |
| D | 9093 hCoV-19/India/TG-CCMB-L1469/2020\|EPI_ISL_471600\|20 | ···················· | ···················· | ···················· | ···················· | ···················· | ···················· |
| D | 9094 hCoV-19/India/TG-CCMB-L1470/2020\|EPI_ISL_471601\|20 | ···················· | ···················· | ···················· | ···················· | ···················· | ···················· |
| D | 9095 hCoV-19/India/TG-CCMB-L1471/2020\|EPI_ISL_471602\|20 | ···················· | ···················· | ···················· | ···················· | ···················· | ···················· |
| D | 9096 hCoV-19/India/TG-CCMB-L1475/2020\|EPI_ISL_471603\|20 | ···················· | ···················· | ···················· | ···················· | ···················· | ···················· |
| D | 9097 hCoV-19/India/TG-CCMB-L1476/2020\|EPI_ISL_471604\|20 | ···················· | ···················· | ···················· | ···················· | ···················· | ···················· |
| D | 9098 hCoV-19/India/TG-CCMB-L1477/2020\|EPI_ISL_471605\|20 | ···················· | ···················· | ···················· | ···················· | ···················· | ···················· |
| D | 9099 hCoV-19/India/TG-CCMB-L1478/2020\|EPI_ISL_471606\|20 | ···················· | ···················· | ···················· | ···················· | ···················· | ···················· |
| D | 9100 hCoV-19/India/TG-CCMB-L1484/2020\|EPI_ISL_471607\|20 | ···················· | ···················· | ···················· | ···················· | ···················· | ···················· |
| D | 9101 hCoV-19/India/TG-CCMB-L1487/2020\|EPI_ISL_471608\|20 | ···················· | ···················· | ···················· | ···················· | ···················· | ···················· |
| D | 9102 hCoV-19/India/TG-CCMB-L1492/2020\|EPI_ISL_471609\|20 | ···················· | ···················· | ···················· | ···················· | ···················· | ···················· |
| D | 9103 hCoV-19/India/TG-CCMB-L1494/2020\|EPI_ISL_471610\|20 | ···················· | ···················· | ···················· | ···················· | ···················· | ···················· |
| D | 9104 hCoV-19/India/TG-CCMB-L1495/2020\|EPI_ISL_471611\|20 | ···················· | ···················· | ···················· | ···················· | ···················· | ···················· |
| D | 9105 hCoV-19/India/TG-CCMB-L1496/2020\|EPI_ISL_471612\|20 | ···················· | ···················· | ···················· | ···················· | ···················· | ···················· |
| D | 9106 hCoV-19/India/TG-CCMB-L1497/2020\|EPI_ISL_471613\|20 | ···················· | ···················· | ···················· | ···················· | ···················· | ···················· |
| D | 9107 hCoV-19/India/TG-CCMB-L1499/2020\|EPI_ISL_471614\|20 | ···················· | ···················· | ···················· | ···················· | ···················· | ···················· |
| D | 9108 hCoV-19/India/TG-CCMB-L1500/2020\|EPI_ISL_471615\|20 | ···················· | ···················· | ···················· | ···················· | ···················· | ···················· |
| D | 9109 hCoV-19/India/TG-CCMB-L1510/2020\|EPI_ISL_471616\|20 | ···················· | ···················· | ···················· | ···················· | ···················· | ···················· |
| D | 9110 hCoV-19/India/TG-CCMB-L1542/2020\|EPI_ISL_471617\|20 | ···················· | ···················· | ···················· | ···················· | ···················· | ···················· |
| D | 9111 hCoV-19/India/TG-CCMB-L1551/2020\|EPI_ISL_471618\|20 | ···················· | ···················· | ···················· | ···················· | ···················· | ···················· |
| D | 9112 hCoV-19/India/TG-CCMB-L1554/2020\|EPI_ISL_471619\|20 | ···················· | ···················· | ···················· | ···················· | ···················· | ···················· |
| D | 9113 hCoV-19/India/TG-CCMB-L1564/2020\|EPI_ISL_471620\|20 | ···················· | ···················· | ···················· | ···················· | ···················· | ···················· |
| D | 9114 hCoV-19/India/TG-CCMB-L1675/2020\|EPI_ISL_471621\|20 | ···················· | ···················· | ···················· | ···················· | ···················· | ···················· |
| D | 9115 hCoV-19/India/TG-CCMB-L1684/2020\|EPI_ISL_471622\|20 | ···················· | ···················· | ···················· | ···················· | ···················· | ···················· |
| D | 9116 hCoV-19/India/TG-CCMB-L1693/2020\|EPI_ISL_471623\|20 | ···················· | ···················· | ···················· | ···················· | ···················· | ···················· |
| D | 9117 hCoV-19/India/TG-CCMB-L1694/2020\|EPI_ISL_471624\|20 | ···················· | ···················· | ···················· | ···················· | ···················· | ···················· |
| D | 9118 hCoV-19/India/TG-CCMB-L1721/2020\|EPI_ISL_471625\|20 | ···················· | ···················· | ···················· | ···················· | ···················· | ···················· |
| D | 9119 hCoV-19/India/TG-CCMB-L1724/2020\|EPI_ISL_471626\|20 | ···················· | ···················· | ···················· | ···················· | ···················· | ···················· |
| D | 9120 hCoV-19/India/TG-CCMB-L1726/2020\|EPI_ISL_471627\|20 | ···················· | ···················· | ···················· | ···················· | ···················· | ···················· |
| D | 9121 hCoV-19/India/TG-CCMB-L1728/2020\|EPI_ISL_471628\|20 | ···················· | ···················· | ···················· | ···················· | ···················· | ···················· |
| D | 9122 hCoV-19/India/TG-CCMB-L1738/2020\|EPI_ISL_471629\|20 | ···················· | ···················· | ···················· | ···················· | ···················· | ···················· |
| D | 9123 hCoV-19/India/TG-CCMB-L1743/2020\|EPI_ISL_471630\|20 | ···················· | ···················· | ···················· | ···················· | ···················· | ···················· |
| D | 9124 hCoV-19/India/TG-CCMB-L1746/2020\|EPI_ISL_471631\|20 | ···················· | ···················· | ···················· | ···················· | ···················· | ···················· |
| D | 9125 hCoV-19/India/TG-CCMB-L1762/2020\|EPI_ISL_471632\|20 | ···················· | ···················· | ···················· | ···················· | ···················· | ···················· |
| D | 9126 hCoV-19/India/TG-CCMB-L1781/2020\|EPI_ISL_471633\|20 | ···················· | ···················· | ···················· | ···················· | ···················· | ···················· |
| D | 9127 hCoV-19/India/TG-CCMB-L1806/2020\|EPI_ISL_471634\|20 | ···················· | ···················· | ···················· | ···················· | ···················· | ···················· |
| D | 9128 hCoV-19/India/TG-CCMB-L1809/2020\|EPI_ISL_471635\|20 | ···················· | ···················· | ···················· | ···················· | ···················· | ···················· |
| D | 9129 hCoV-19/India/TG-CCMB-L1810/2020\|EPI_ISL_471636\|20 | ···················· | ···················· | ···················· | ···················· | ···················· | ···················· |
| D | 9130 hCoV-19/India/TG-CCMB-L1811/2020\|EPI_ISL_471637\|20 | ···················· | ···················· | ···················· | ···················· | ···················· | ···················· |
| D | 9131 hCoV-19/India/TG-CCMB-L1819/2020\|EPI_ISL_471638\|20 | ···················· | ···················· | ···················· | ···················· | ···················· | ···················· |
| D | 9132 hCoV-19/India/TG-CCMB-L1822/2020\|EPI_ISL_471639\|20 | ···················· | ···················· | ···················· | ···················· | ···················· | ···················· |
| D | 9133 hCoV-19/India/TG-CCMB-L1824/2020\|EPI_ISL_471640\|20 | ···················· | ···················· | ···················· | ···················· | ···················· | ···················· |
| D | 9134 hCoV-19/India/TG-CCMB-L276-P3/2020\|EPI_ISL_471641\| | ···················· | ···················· | ···················· | ···················· | ···················· | ···················· |
| D | 9135 hCoV-19/India/TG-CCMB-L301-P3/2020\|EPI_ISL_471642\| | ···················· | ···················· | ···················· | ···················· | ···················· | ···················· |
| D | 9136 hCoV-19/India/TG-CCMB-L302/2020\|EPI_ISL_471643\|202 | ···················· | ···················· | ···················· | ···················· | ···················· | ···················· |
| D | 9137 hCoV-19/India/TG-CCMB-L912/2020\|EPI_ISL_471644\|202 | ···················· | ···················· | ···················· | ···················· | ···················· | ···················· |
| D | 9138 hCoV-19/India/TG-CCMB-O2-P4M/2020\|EPI_ISL_471645\|2 | ···················· | ···················· | ···················· | ···················· | ···················· | ···················· |
| D | 9139 hCoV-19/India/TG-CCMB-O2-P4S/2020\|EPI_ISL_471646\|2 | ···················· | ···················· | ···················· | ···················· | ···················· | ···················· |
| D | 9140 hCoV-19/Brazil/SP-526/2020\|EPI_ISL_471647\|2020-04- | ···················· | ···················· | ···················· | ···················· | ···················· | ···················· |
| D | 9141 hCoV-19/Brazil/SP-527/2020\|EPI_ISL_471648\|2020-04- | ···················· | ···················· | ···················· | ···················· | ···················· | ···················· |
| D | 9142 hCoV-19/USA/MI-MDHHS-SC20655/2020\|EPI_ISL_471679\|2 | ···················· | ···················· | ···················· | ···················· | ···················· | ···················· |
| D | 9143 hCoV-19/USA/MI-MDHHS-SC20661/2020\|EPI_ISL_471685\|2 | ···················· | ···················· | ···················· | ···················· | ···················· | ···················· |
| D | 9144 hCoV-19/USA/MI-MDHHS-SC20662/2020\|EPI_ISL_471686\|2 | ···················· | ···················· | ···················· | ···················· | ···················· | ···················· |
| D | 9145 hCoV-19/USA/MI-MDHHS-SC20663/2020\|EPI_ISL_471687\|2 | ···················· | ···················· | ···················· | ···················· | ···················· | ···················· |
| D | 9146 hCoV-19/USA/MI-MDHHS-SC20665/2020\|EPI_ISL_471689\|2 | ···················· | ···················· | ···················· | ···················· | ···················· | ···················· |
| D | 9147 hCoV-19/USA/MI-MDHHS-SC20667/2020\|EPI_ISL_471691\|2 | ···················· | ···················· | ···················· | ···················· | ···················· | ···················· |
| D | 9148 hCoV-19/USA/MI-MDHHS-SC20668/2020\|EPI_ISL_471692\|2 | ···················· | ···················· | ···················· | ···················· | ···················· | ···················· |
| D | 9149 hCoV-19/USA/MI-MDHHS-SC20669/2020\|EPI_ISL_471693\|2 | ···················· | ···················· | ···················· | ···················· | ···················· | ···················· |
| D | 9150 hCoV-19/USA/MI-MDHHS-SC20671/2020\|EPI_ISL_471695\|2 | ···················· | ···················· | ···················· | ···················· | ···················· | ···················· |
| D | 9151 hCoV-19/USA/MI-MDHHS-SC20672/2020\|EPI_ISL_471696\|2 | ···················· | ···················· | ···················· | ···················· | ···················· | ···················· |
| D | 9152 hCoV-19/Wales/PHWC-160270/2020\|EPI_ISL_472913\|2020 | ···················· | ···················· | ···················· | ···················· | ···················· | ···················· |
| D | 9153 hCoV-19/Wales/PHWC-16029E/2020\|EPI_ISL_472914\|2020 | ···················· | ···················· | ···················· | ···················· | ···················· | ···················· |
| D | 9154 hCoV-19/Wales/PHWC-1602CB/2020\|EPI_ISL_472916\|2020 | ···················· | ···················· | ···················· | ···················· | ···················· | ···················· |
| D | 9155 hCoV-19/Wales/PHWC-16035F/2020\|EPI_ISL_472922\|2020 | ···················· | ···················· | ···················· | ···················· | ···················· | ···················· |
| D | 9156 hCoV-19/Wales/PHWC-1603B9/2020\|EPI_ISL_472926\|2020 | ···················· | ···················· | ···················· | ···················· | ···················· | ···················· |
| D | 9157 hCoV-19/Wales/PHWC-1603C8/2020\|EPI_ISL_472927\|2020 | ···················· | ···················· | ···················· | ···················· | ···················· | ···················· |
| D | 9158 hCoV-19/Wales/PHWC-16044D/2020\|EPI_ISL_472934\|2020 | ···················· | ····A··············· | ···················· | ···················· | ···················· | ···················· |

| | | | | | | | | |
|---|---|---|---|---|---|---|---|---|
| D | 9159 | hCoV-19/Wales/PHWC-16045C/2020\|EPI_ISL_472935\|2020 | ···················· | ···················· | ···················· | ···················· | ···················· | ···················· |
| D | 9160 | hCoV-19/Wales/PHWC-160489/2020\|EPI_ISL_472937\|2020 | ···················· | ···················· | ···················· | ···················· | ···················· | ···················· |
| D | 9161 | hCoV-19/Wales/PHWC-160498/2020\|EPI_ISL_472938\|2020 | ···················· | ···················· | ···················· | ···················· | ···················· | ···················· |
| D | 9162 | hCoV-19/Wales/PHWC-1604A7/2020\|EPI_ISL_472939\|2020 | ···················· | ···················· | ···················· | ···················· | ···················· | ···················· |
| D | 9163 | hCoV-19/Wales/PHWC-1604B6/2020\|EPI_ISL_472940\|2020 | ···················· | ···················· | ···················· | ···················· | ···················· | ···················· |
| D | 9164 | hCoV-19/Wales/PHWC-1604D4/2020\|EPI_ISL_472942\|2020 | ···················· | ···················· | ···················· | ···················· | ···················· | ···················· |
| D | 9165 | hCoV-19/Wales/PHWC-16052C/2020\|EPI_ISL_472947\|2020 | ···················· | ···················· | ···················· | ···················· | ···················· | ···················· |
| D | 9166 | hCoV-19/Wales/PHWC-16054A/2020\|EPI_ISL_472948\|2020 | ···················· | ···················· | ···················· | ···················· | ···················· | ···················· |
| D | 9167 | hCoV-19/Wales/PHWC-160559/2020\|EPI_ISL_472949\|2020 | ···················· | ···················· | ···················· | ···················· | ···················· | ···················· |
| D | 9168 | hCoV-19/Wales/PHWC-1605C2/2020\|EPI_ISL_472952\|2020 | ···················· | ···················· | ···················· | ···················· | ···················· | ···················· |
| D | 9169 | hCoV-19/Wales/PHWC-160629/2020\|EPI_ISL_472955\|2020 | ···················· | ···················· | ···················· | ···················· | ···················· | ···················· |
| D | 9170 | hCoV-19/Wales/PHWC-160638/2020\|EPI_ISL_472956\|2020 | ···················· | ···················· | ···················· | ···················· | ···················· | ···················· |
| D | 9171 | hCoV-19/Wales/PHWC-160647/2020\|EPI_ISL_472957\|2020 | ···················· | ····A··············· | ···················· | ···················· | ···················· | ···················· |
| D | 9172 | hCoV-19/Wales/PHWC-160674/2020\|EPI_ISL_472960\|2020 | ···················· | ···················· | ···················· | ···················· | ···················· | ···················· |
| D | 9173 | hCoV-19/Wales/PHWC-160683/2020\|EPI_ISL_472961\|2020 | ···················· | ···················· | ···················· | ···················· | ···················· | ···················· |
| D | 9174 | hCoV-19/Wales/PHWC-1606A1/2020\|EPI_ISL_472963\|2020 | ···················· | ···················· | ···················· | ···················· | ···················· | ···················· |
| D | 9175 | hCoV-19/Wales/PHWC-1606CF/2020\|EPI_ISL_472965\|2020 | ···················· | ···················· | ···················· | ···················· | ···················· | ···················· |
| D | 9176 | hCoV-19/Wales/PHWC-1606DE/2020\|EPI_ISL_472966\|2020 | ···················· | ···················· | ···················· | ···················· | ···················· | ···················· |
| D | 9177 | hCoV-19/Wales/PHWC-1606ED/2020\|EPI_ISL_472967\|2020 | ···················· | ···················· | ···················· | ···················· | ···················· | ···················· |
| D | 9178 | hCoV-19/Wales/PHWC-160708/2020\|EPI_ISL_472968\|2020 | ···················· | ···················· | ···················· | ···················· | ···················· | ···················· |
| D | 9179 | hCoV-19/Wales/PHWC-160744/2020\|EPI_ISL_472972\|2020 | ···················· | ···················· | ···················· | ···················· | ···················· | ···················· |
| D | 9181 | hCoV-19/Wales/PHWC-160780/2020\|EPI_ISL_472975\|2020 | ···················· | ···················· | ···················· | ···················· | ···················· | ···················· |
| D | 9182 | hCoV-19/Wales/PHWC-16079F/2020\|EPI_ISL_472976\|2020 | ···················· | ···················· | ···················· | ···················· | ···················· | ···················· |
| D | 9183 | hCoV-19/Wales/PHWC-1607F9/2020\|EPI_ISL_472979\|2020 | ···················· | ···················· | ···················· | ···················· | ···················· | ···················· |
| D | 9184 | hCoV-19/Wales/PHWC-160805/2020\|EPI_ISL_472980\|2020 | ···················· | ···················· | ···················· | ···················· | ···················· | ···················· |
| D | 9185 | hCoV-19/Wales/PHWC-160832/2020\|EPI_ISL_472983\|2020 | ···················· | ···················· | ···················· | ···················· | ···················· | ···················· |
| D | 9186 | hCoV-19/Wales/PHWC-1608AB/2020\|EPI_ISL_472988\|2020 | ···················· | ···················· | ···················· | ···················· | ···················· | ···················· |
| D | 9187 | hCoV-19/Wales/PHWC-1608C9/2020\|EPI_ISL_472990\|2020 | ···················· | ···················· | ···················· | ···················· | ···················· | ···················· |
| D | 9188 | hCoV-19/Wales/PHWC-1608D8/2020\|EPI_ISL_472991\|2020 | ···················· | ···················· | ···················· | ···················· | ···················· | ···················· |
| D | 9189 | hCoV-19/Wales/PHWC-160920/2020\|EPI_ISL_472994\|2020 | ···················· | ···················· | ···················· | ···················· | ···················· | ···················· |
| D | 9190 | hCoV-19/Wales/PHWC-16093F/2020\|EPI_ISL_472995\|2020 | ···················· | ···················· | ···················· | ···················· | ···················· | ···················· |
| D | 9191 | hCoV-19/Wales/PHWC-16095D/2020\|EPI_ISL_472997\|2020 | ···················· | ···················· | ···················· | ···················· | ···················· | ···················· |
| D | 9192 | hCoV-19/Wales/PHWC-16096C/2020\|EPI_ISL_472998\|2020 | ···················· | ···················· | ···················· | ···················· | ···················· | ···················· |
| D | 9193 | hCoV-19/Wales/PHWC-163C24/2020\|EPI_ISL_474058\|2020 | ···················· | ···················· | ···················· | ···················· | ···················· | ···················· |
| D | 9194 | hCoV-19/Wales/PHWC-163C33/2020\|EPI_ISL_474059\|2020 | ···················· | ···················· | ···················· | ···················· | ···················· | ···················· |
| D | 9195 | hCoV-19/Wales/PHWC-163C42/2020\|EPI_ISL_474060\|2020 | ···················· | ···················· | ···················· | ···················· | ···················· | ···················· |
| D | 9196 | hCoV-19/Wales/PHWC-163C51/2020\|EPI_ISL_474061\|2020 | ···················· | ···················· | ···················· | ···················· | ···················· | ···················· |
| D | 9197 | hCoV-19/Wales/PHWC-163C60/2020\|EPI_ISL_474062\|2020 | ···················· | ···················· | ···················· | ···················· | ···················· | ···················· |
| D | 9199 | hCoV-19/Wales/PHWC-163C9D/2020\|EPI_ISL_474065\|2020 | ···················· | ···················· | ···················· | ···················· | ···················· | ···················· |
| D | 9200 | hCoV-19/Wales/PHWC-163CBB/2020\|EPI_ISL_474067\|2020 | ···················· | ····A··············· | ···················· | ···················· | ···················· | ···················· |
| D | 9201 | hCoV-19/Wales/PHWC-163CE8/2020\|EPI_ISL_474069\|2020 | ···················· | ···················· | ···················· | ···················· | ···················· | ···················· |
| D | 9202 | hCoV-19/Wales/PHWC-163D03/2020\|EPI_ISL_474071\|2020 | ···················· | ···················· | ···················· | ···················· | ···················· | ···················· |
| D | 9203 | hCoV-19/Wales/PHWC-163D21/2020\|EPI_ISL_474073\|2020 | ···················· | ···················· | ···················· | ···················· | ···················· | ···················· |
| D | 9204 | hCoV-19/Wales/PHWC-163D6D/2020\|EPI_ISL_474077\|2020 | ···················· | ···················· | ···················· | ···················· | ···················· | ···················· |
| D | 9205 | hCoV-19/Wales/PHWC-163D7C/2020\|EPI_ISL_474078\|2020 | ···················· | ···················· | ···················· | ···················· | ···················· | ···················· |
| D | 9207 | hCoV-19/USA/NE-10862/2020\|EPI_ISL_475191\|2020-05-0 | ···················· | ···················· | ···················· | ···················· | ···················· | ···················· |
| D | 9208 | hCoV-19/USA/NE-10866/2020\|EPI_ISL_475192\|2020-05-0 | ···················· | ···················· | ···················· | ···················· | ···················· | ···················· |
| D | 9209 | hCoV-19/USA/NE-10889/2020\|EPI_ISL_475194\|2020-05-0 | ···················· | ···················· | ···················· | ···················· | ···················· | ···················· |
| D | 9210 | hCoV-19/USA/NE-10943/2020\|EPI_ISL_475195\|2020-05-0 | ···················· | ···················· | ···················· | ···················· | ···················· | ···················· |
| D | 9211 | hCoV-19/USA/NE-10964/2020\|EPI_ISL_475196\|2020-05-0 | ···················· | ···················· | ···················· | ···················· | ···················· | ···················· |
| D | 9212 | hCoV-19/USA/NE-10970/2020\|EPI_ISL_475197\|2020-05-0 | ···················· | ···················· | ···················· | ···················· | ···················· | ···················· |
| D | 9213 | hCoV-19/USA/NE-11111/2020\|EPI_ISL_475198\|2020-05-0 | ···················· | ···················· | ···················· | ···················· | ···················· | ···················· |
| D | 9214 | hCoV-19/England/SHEF-C5E67/2020\|EPI_ISL_475389\|202 | ···················· | ···················· | ···················· | ···················· | ···················· | ···················· |
| D | 9215 | hCoV-19/England/SHEF-C630D/2020\|EPI_ISL_475391\|202 | ···················· | ···················· | ···················· | ···················· | ···················· | ···················· |
| D | 9216 | hCoV-19/England/SHEF-C6B41/2020\|EPI_ISL_475393\|202 | ···················· | ···················· | ···················· | ···················· | ···················· | ···················· |
| D | 9217 | hCoV-19/Bangladesh/BCSIR-NILMRC-004/2020\|EPI_ISL_4 | ···················· | ···················· | ···················· | ···················· | ···················· | ···················· |
| D | 9218 | hCoV-19/Bangladesh/BCSIR-NILMRC-002/2020\|EPI_ISL_4 | ···················· | ···················· | ···················· | ···················· | ···················· | ···················· |
| D | 9219 | hCoV-19/Bangladesh/BCSIR-NILMRC-003/2020\|EPI_ISL_4 | ···················· | ···················· | ···················· | ···················· | ···················· | ···················· |
| D | 9220 | hCoV-19/Bangladesh/BCSIR-NILMRC-005/2020\|EPI_ISL_4 | ···················· | ···················· | ···················· | ···················· | ···················· | ···················· |
| D | 9221 | hCoV-19/Bangladesh/BCSIR-NILMRC-006.2/2020\|EPI_ISL | ···················· | ···················· | ···················· | ···················· | ···················· | ···················· |
| D | 9222 | hCoV-19/Bangladesh/BCSIR-NILMRC-009/2020\|EPI_ISL_4 | ···················· | ···················· | ···················· | ···················· | ···················· | ···················· |
| D | 9223 | hCoV-19/Bangladesh/BCSIR-NILMRC-015/2020\|EPI_ISL_4 | ···················· | ···················· | ···················· | ···················· | ···················· | ···················· |
| D | 9224 | hCoV-19/Bangladesh/BCSIR-NILMRC-042/2020\|EPI_ISL_4 | ···················· | ···················· | ···················· | ···················· | ···················· | ···················· |
| D | 9225 | hCoV-19/Bangladesh/BCSIR-NILMRC-050/2020\|EPI_ISL_4 | ···················· | ···················· | ···················· | ···················· | ···················· | ···················· |
| D | 9226 | hCoV-19/USA/LA-BIE-098/2020\|EPI_ISL_463300\|2020-04 | ···················· | ···················· | ···················· | ···················· | ···················· | ···················· |
| D | 9227 | hCoV-19/USA/WA-S835/2020\|EPI_ISL_463325\|2020-04-21 | ···················· | ···················· | ···················· | ···················· | ···················· | ···················· |
| D | 9228 | hCoV-19/USA/WA-S836/2020\|EPI_ISL_463326\|2020-04-21 | ···················· | ···················· | ···················· | ···················· | ···················· | ···················· |
| D | 9229 | hCoV-19/USA/WA-S837/2020\|EPI_ISL_463327\|2020-04-20 | ···················· | ···················· | ···················· | ···················· | ···················· | ···················· |
| D | 9230 | hCoV-19/USA/WA-S838/2020\|EPI_ISL_463328\|2020-04-22 | ···················· | ···················· | ···················· | ···················· | ···················· | ···················· |
| D | 9231 | hCoV-19/USA/WA-S839/2020\|EPI_ISL_463329\|2020-04-21 | ···················· | ···················· | ···················· | ···················· | ···················· | ···················· |
| D | 9232 | hCoV-19/USA/WA-S840/2020\|EPI_ISL_463330\|2020-04-18 | ···················· | ···················· | ···················· | ···················· | ···················· | ···················· |
| D | 9233 | hCoV-19/USA/WA-S841/2020\|EPI_ISL_463331\|2020-04-21 | ···················· | ···················· | ···················· | ···················· | ···················· | ···················· |
| D | 9234 | hCoV-19/USA/WA-S842/2020\|EPI_ISL_463332\|2020-04-21 | ···················· | ···················· | ···················· | ···················· | ···················· | ···················· |
| D | 9235 | hCoV-19/USA/WA-S843/2020\|EPI_ISL_463333\|2020-04-22 | ···················· | ···················· | ···················· | ···················· | ···················· | ···················· |
| D | 9236 | hCoV-19/USA/WA-S844/2020\|EPI_ISL_463334\|2020-04-22 | ···················· | ···················· | ···················· | ···················· | ···················· | ···················· |
| D | 9237 | hCoV-19/USA/WA-S845/2020\|EPI_ISL_463335\|2020-04-23 | ···················· | N·····N············· | ···················· | ···················· | ···················· | ···················· |
| D | 9238 | hCoV-19/USA/WA-S847/2020\|EPI_ISL_463337\|2020-04-23 | ···················· | ···················· | ···················· | ···················· | ···················· | ···················· |
| D | 9239 | hCoV-19/USA/WA-S848/2020\|EPI_ISL_463338\|2020-04-23 | ···················· | ···················· | ···················· | ···················· | ···················· | ···················· |

| | | | | | | | | |
|---|---|---|---|---|---|---|---|---|
| D | 9240 hCoV-19/USA/WA-S849/2020\|EPI_ISL_463339\|2020-04-23 | ···················· | ···················· | ···················· | ···················· | ···················· | ···················· | |
| D | 9241 hCoV-19/USA/WA-S850/2020\|EPI_ISL_463340\|2020-04-23 | ···················· | ···················· | ···················· | ···················· | ···················· | ···················· | |
| D | 9242 hCoV-19/USA/WA-S851/2020\|EPI_ISL_463341\|2020-04-24 | ···················· | ···················· | ···················· | ···················· | ···················· | ···················· | |
| D | 9243 hCoV-19/USA/WA-S852/2020\|EPI_ISL_463342\|2020-04-24 | ···················· | ···················· | ···················· | ···················· | ···················· | ···················· | |
| D | 9244 hCoV-19/USA/WA-S853/2020\|EPI_ISL_463343\|2020-04-24 | ···················· | ···················· | ···················· | ···················· | ···················· | ···················· | |
| D | 9245 hCoV-19/USA/WA-S854/2020\|EPI_ISL_463344\|2020-04-24 | ···················· | ···················· | ···················· | ···················· | ···················· | ···················· | |
| D | 9246 hCoV-19/USA/WA-S856/2020\|EPI_ISL_463346\|2020-04-24 | ···················· | ···················· | ···················· | ···················· | ···················· | ···················· | |
| D | 9247 hCoV-19/USA/WA-S857/2020\|EPI_ISL_463347\|2020-04-24 | ···················· | ···················· | ···················· | ···················· | ···················· | ···················· | |
| D | 9248 hCoV-19/USA/WA-S858/2020\|EPI_ISL_463348\|2020-04-24 | ···················· | ···················· | ···················· | ···················· | ···················· | ···················· | |
| D | 9249 hCoV-19/USA/WA-S859/2020\|EPI_ISL_463349\|2020-04-25 | ···················· | ···················· | ···················· | ···················· | ···················· | ···················· | |
| D | 9250 hCoV-19/USA/WA-S860/2020\|EPI_ISL_463350\|2020-04-24 | ···················· | ···················· | ···················· | ···················· | ···················· | ···················· | |
| D | 9251 hCoV-19/USA/WA-S861/2020\|EPI_ISL_463351\|2020-04-23 | ···················· | ·A·················· | ···················· | ···················· | ···················· | ···················· | |
| D | 9252 hCoV-19/USA/WA-S862/2020\|EPI_ISL_463352\|2020-04-22 | ···················· | ···················· | ···················· | ···················· | ···················· | ···················· | |
| D | 9253 hCoV-19/USA/WA-S863/2020\|EPI_ISL_463353\|2020-04-22 | ···················· | ···················· | ···················· | ···················· | ···················· | ···················· | |
| D | 9254 hCoV-19/USA/WA-S864/2020\|EPI_ISL_463354\|2020-04-23 | ···················· | ···················· | ···················· | ···················· | ···················· | ···················· | |
| D | 9255 hCoV-19/USA/WA-S865/2020\|EPI_ISL_463355\|2020-04-22 | ···················· | ···················· | ···················· | ···················· | ···················· | ···················· | |
| D | 9256 hCoV-19/USA/WA-S866/2020\|EPI_ISL_463356\|2020-04-24 | ···················· | ···················· | ···················· | ···················· | ···················· | ···················· | |
| D | 9257 hCoV-19/USA/WA-S867/2020\|EPI_ISL_463357\|2020-04-24 | ···················· | ···················· | ···················· | ···················· | ···················· | ···················· | |
| D | 9258 hCoV-19/USA/WA-S868/2020\|EPI_ISL_463358\|2020-04-23 | ···················· | ···················· | ···················· | ···················· | ···················· | ···················· | |
| D | 9259 hCoV-19/USA/WA-S869/2020\|EPI_ISL_463359\|2020-04-23 | ···················· | ···················· | ···················· | ···················· | ···················· | ···················· | |
| D | 9260 hCoV-19/USA/WA-S870/2020\|EPI_ISL_463360\|2020-04-24 | ···················· | ···················· | ···················· | ···················· | ···················· | ···················· | |
| D | 9261 hCoV-19/USA/WA-S871/2020\|EPI_ISL_463361\|2020-04-24 | ···················· | ···················· | ···················· | ···················· | ···················· | ···················· | |
| D | 9262 hCoV-19/USA/WA-S872/2020\|EPI_ISL_463362\|2020-04-24 | ···················· | ···················· | ···················· | ···················· | ···················· | ···················· | |
| D | 9263 hCoV-19/USA/WA-S873/2020\|EPI_ISL_463363\|2020-04-16 | ···················· | ···················· | ···················· | ···················· | ···················· | ···················· | |
| D | 9264 hCoV-19/USA/WA-S874/2020\|EPI_ISL_463364\|2020-04-16 | ···················· | ···················· | ···················· | ···················· | ···················· | ···················· | |
| D | 9265 hCoV-19/USA/WA-S875/2020\|EPI_ISL_463365\|2020-04-16 | ···················· | ···················· | ···················· | ···················· | ···················· | ···················· | |
| D | 9266 hCoV-19/USA/WA-S876/2020\|EPI_ISL_463366\|2020-04-16 | ···················· | ···················· | ···················· | ···················· | ···················· | ···················· | |
| D | 9267 hCoV-19/USA/WA-S877/2020\|EPI_ISL_463367\|2020-04-15 | ···················· | ···················· | ···················· | ···················· | ···················· | ···················· | |
| D | 9268 hCoV-19/USA/WA-S879/2020\|EPI_ISL_463369\|2020-04-23 | ···················· | ···················· | ···················· | ···················· | ···················· | ···················· | |
| D | 9269 hCoV-19/USA/WA-S880/2020\|EPI_ISL_463370\|2020-04-23 | ···················· | ···················· | ···················· | ···················· | ···················· | ···················· | |
| D | 9270 hCoV-19/USA/WA-S881/2020\|EPI_ISL_463371\|2020-04-23 | ···················· | ···················· | ···················· | ···················· | ···················· | ···················· | |
| D | 9271 hCoV-19/USA/WA-S888/2020\|EPI_ISL_463378\|2020-04-22 | ···················· | ···················· | ···················· | ···················· | ···················· | ···················· | |
| D | 9272 hCoV-19/USA/WA-S889/2020\|EPI_ISL_463379\|2020-04-28 | ···················· | ···················· | ···················· | ···················· | ···················· | ···················· | |
| D | 9273 hCoV-19/USA/WA-S891/2020\|EPI_ISL_463381\|2020-04-28 | ···················· | ···················· | ···················· | ···················· | ···················· | ···················· | |
| D | 9274 hCoV-19/USA/WA-S892/2020\|EPI_ISL_463382\|2020-04-28 | ···················· | ···················· | ···················· | ···················· | ···················· | ···················· | |
| D | 9275 hCoV-19/USA/WA-S893/2020\|EPI_ISL_463383\|2020-04-28 | ···················· | ···················· | ···················· | ···················· | ···················· | ···················· | |
| D | 9276 hCoV-19/USA/WA-S895/2020\|EPI_ISL_463385\|2020-04-24 | ···················· | ···················· | ···················· | ···················· | ···················· | ···················· | |
| D | 9277 hCoV-19/USA/WA-S896/2020\|EPI_ISL_463386\|2020-04-28 | ···················· | ···················· | ···················· | ···················· | ···················· | ···················· | |
| D | 9278 hCoV-19/England/201090978/2020\|EPI_ISL_464614\|2020 | ···················· | ···················· | ···················· | ···················· | ···················· | ···················· | |
| D | 9281 hCoV-19/England/201101382/2020\|EPI_ISL_464660\|2020 | ···················· | ···················· | ···················· | ···················· | ···················· | ···················· | |
| D | 9283 hCoV-19/England/201120242/2020\|EPI_ISL_464672\|2020 | ···················· | ···················· | ···················· | ···················· | ···················· | ···················· | |
| D | 9284 hCoV-19/England/201121119/2020\|EPI_ISL_464682\|2020 | ···················· | ···················· | ···················· | ···················· | ···················· | ···················· | |
| D | 9285 hCoV-19/England/201121123/2020\|EPI_ISL_464684\|2020 | ···················· | ···················· | ···················· | ···················· | ··NNN··············· | ···················· | 1 |
| D | 9286 hCoV-19/England/20142027104/2020\|EPI_ISL_465940\|20 | ···················· | ···················· | ···················· | ···················· | ···················· | ···················· | |
| D | 9287 hCoV-19/England/20142027704/2020\|EPI_ISL_465941\|20 | ···················· | ···················· | ···················· | ···················· | ···················· | ···················· | |
| D | 9288 hCoV-19/England/20142078104/2020\|EPI_ISL_465943\|20 | ···················· | ···················· | ···················· | ···················· | ···················· | ···················· | |
| D | 9289 hCoV-19/England/20142078204/2020\|EPI_ISL_465944\|20 | ···················· | ···················· | ···················· | ···················· | ···················· | ···················· | |
| D | 9290 hCoV-19/England/20142084004/2020\|EPI_ISL_465946\|20 | ···················· | ···················· | ···················· | ···················· | ···················· | ···················· | |
| D | 9291 hCoV-19/England/20142130904/2020\|EPI_ISL_465960\|20 | ···················· | ···················· | ···················· | ···················· | ···················· | ···················· | |
| D | 9292 hCoV-19/England/20142131104/2020\|EPI_ISL_465962\|20 | ···················· | ···················· | ···················· | ···················· | ···················· | ···················· | |
| D | 9294 hCoV-19/England/20144000104/2020\|EPI_ISL_465971\|20 | ···················· | ···················· | ···················· | ···················· | ···················· | ···················· | |
| D | 9295 hCoV-19/England/20144003904/2020\|EPI_ISL_465973\|20 | ···················· | ···················· | ···················· | ···················· | ···················· | ···················· | |
| D | 9296 hCoV-19/England/20144009404/2020\|EPI_ISL_465975\|20 | ···················· | ···················· | ···················· | ···················· | ···················· | ···················· | |
| D | 9297 hCoV-19/England/20144011304/2020\|EPI_ISL_465976\|20 | ···················· | ···················· | ···················· | ···················· | ···················· | ···················· | |
| D | 9298 hCoV-19/England/20144011404/2020\|EPI_ISL_465977\|20 | ···················· | ···················· | ···················· | ···················· | ···················· | ···················· | |
| D | 9299 hCoV-19/England/20144012104/2020\|EPI_ISL_465978\|20 | ···················· | ···················· | ···················· | ···················· | ···················· | ···················· | |
| D | 9300 hCoV-19/England/201440125/2020\|EPI_ISL_465980\|2020 | ···················· | ···················· | ···················· | ···················· | ···················· | ···················· | |
| D | 9301 hCoV-19/England/201440167/2020\|EPI_ISL_465984\|2020 | ···················· | ···················· | ···················· | ···················· | ···················· | ···················· | |
| D | 9302 hCoV-19/England/20144021704/2020\|EPI_ISL_465987\|20 | ···················· | ···················· | ···················· | ···················· | ···················· | ···················· | |
| D | 9303 hCoV-19/England/20144022004/2020\|EPI_ISL_465990\|20 | ···················· | ···················· | ···················· | ··M················· | ···················· | ···················· | |
| D | 9304 hCoV-19/England/20144022204/2020\|EPI_ISL_465991\|20 | ···················· | ···················· | ···················· | ···················· | ···················· | ···················· | |
| D | 9305 hCoV-19/England/20144022604/2020\|EPI_ISL_465992\|20 | ···················· | ···················· | ···················· | ···················· | ···················· | ···················· | |
| D | 9306 hCoV-19/England/20144023004/2020\|EPI_ISL_465993\|20 | ···················· | ···················· | ···················· | ···················· | ···················· | ···················· | |
| D | 9307 hCoV-19/England/20144023304/2020\|EPI_ISL_465994\|20 | ···················· | ···················· | ···················· | ···················· | ···················· | ···················· | |
| D | 9308 hCoV-19/England/20144023504/2020\|EPI_ISL_465995\|20 | ···················· | ···················· | ···················· | ···················· | ···················· | ···················· | |
| D | 9309 hCoV-19/England/20144023604/2020\|EPI_ISL_465996\|20 | ···················· | ···················· | ···················· | ···················· | ···················· | ···················· | |
| D | 9310 hCoV-19/England/20144024004/2020\|EPI_ISL_465997\|20 | ···················· | ·A·················· | ···················· | ···················· | ···················· | ···················· | |
| D | 9311 hCoV-19/England/20144037104/2020\|EPI_ISL_465999\|20 | ···················· | ···················· | ···················· | ···················· | ···················· | ···················· | |
| D | 9312 hCoV-19/England/BRIS-12C841/2020\|EPI_ISL_469500\|20 | ···················· | ···················· | ···················· | ···················· | ···················· | ···················· | |
| D | 9313 hCoV-19/England/BRIS-12C762/2020\|EPI_ISL_469502\|20 | ···················· | ···················· | ···················· | ···················· | ···················· | ···················· | |
| D | 9314 hCoV-19/England/BRIS-12C9D5/2020\|EPI_ISL_469503\|20 | ···················· | ···················· | ···················· | ···················· | ···················· | ···················· | |
| D | 9315 hCoV-19/USA/MI-MDHHS-SC20786/2020\|EPI_ISL_471807\|2 | ···················· | ···················· | ···················· | ···················· | ···················· | ···················· | |
| D | 9316 hCoV-19/USA/MI-MDHHS-SC20787/2020\|EPI_ISL_471808\|2 | ···················· | ···················· | ···················· | ···················· | ···················· | ···················· | |
| D | 9317 hCoV-19/USA/MI-MDHHS-SC20788/2020\|EPI_ISL_471809\|2 | ···················· | ···················· | ···················· | ···················· | ···················· | ···················· | |
| D | 9318 hCoV-19/USA/MI-MDHHS-SC20789/2020\|EPI_ISL_471810\|2 | ···················· | ···················· | ···················· | ···················· | ···················· | ···················· | |
| D | 9319 hCoV-19/USA/MI-MDHHS-SC20791/2020\|EPI_ISL_471812\|2 | ···················· | ···················· | ···················· | ···················· | ···················· | ···················· | |
| D | 9320 hCoV-19/USA/MI-MDHHS-SC20792/2020\|EPI_ISL_471813\|2 | ···················· | ···················· | ···················· | ···················· | ···················· | ···················· | |
| D | 9321 hCoV-19/USA/MI-MDHHS-SC20793/2020\|EPI_ISL_471814\|2 | ···················· | ···················· | ···················· | ···················· | ···················· | ···················· | |

| | | | | | | | |
|---|---|---|---|---|---|---|---|
| D | 9322 hCoV-19/USA/MI-MDHHS-SC20794/2020\|EPI_ISL_471815\|2 | .................... | .................... | ..................... | .................... | .................... | .................... |
| D | 9323 hCoV-19/USA/MI-MDHHS-SC20796/2020\|EPI_ISL_471817\|2 | .................... | .................... | ..................... | .................... | .................... | .................... |
| D | 9324 hCoV-19/USA/MI-MDHHS-SC20797/2020\|EPI_ISL_471818\|2 | .................... | .................... | ..................... | .................... | .................... | .................... |
| D | 9325 hCoV-19/USA/MI-MDHHS-SC20798/2020\|EPI_ISL_471819\|2 | .................... | .................... | ..................... | .................... | .................... | .................... |
| D | 9326 hCoV-19/Wales/PHWC-163802/2020\|EPI_ISL_474000\|2020 | .................... | .................... | ..................... | .................... | .................... | .................... |
| D | 9327 hCoV-19/Wales/PHWC-163811/2020\|EPI_ISL_474001\|2020 | .................... | ....A............... | ..................... | .................... | .................... | .................... |
| D | 9328 hCoV-19/Wales/PHWC-16386C/2020\|EPI_ISL_474006\|2020 | .................... | .................... | ..................... | .................... | .................... | .................... |
| D | 9329 hCoV-19/Wales/PHWC-16387B/2020\|EPI_ISL_474007\|2020 | .................... | .................... | ..................... | .................... | .................... | .................... |
| D | 9330 hCoV-19/Wales/PHWC-163899/2020\|EPI_ISL_474009\|2020 | .................... | .................... | ..................... | .................... | .................... | .................... |
| D | 9331 hCoV-19/Wales/PHWC-1638A8/2020\|EPI_ISL_474010\|2020 | .................... | .................... | ..................... | .................... | .................... | .................... |
| D | 9332 hCoV-19/Wales/PHWC-1638B7/2020\|EPI_ISL_474011\|2020 | .................... | .................... | ..................... | .................... | .................... | .................... |
| D | 9333 hCoV-19/Wales/PHWC-1638C6/2020\|EPI_ISL_474012\|2020 | .................... | .................... | ..................... | .................... | .................... | .................... |
| D | 9334 hCoV-19/Wales/PHWC-1638D5/2020\|EPI_ISL_474013\|2020 | .................... | ....A............... | ..................... | .................... | .................... | .................... |
| D | 9335 hCoV-19/Wales/PHWC-163969/2020\|EPI_ISL_474020\|2020 | .................... | .................... | ..................... | .................... | .................... | .................... |
| D | 9336 hCoV-19/Wales/PHWC-163978/2020\|EPI_ISL_474021\|2020 | .................... | .................... | ..................... | .................... | .................... | .................... |
| D | 9337 hCoV-19/Wales/PHWC-163996/2020\|EPI_ISL_474022\|2020 | .................... | .................... | ..................... | .................... | .................... | .................... |
| D | 9338 hCoV-19/Wales/PHWC-1639F0/2020\|EPI_ISL_474026\|2020 | .................... | .................... | ..................... | .................... | .................... | .................... |
| D | 9339 hCoV-19/Wales/PHWC-163A39/2020\|EPI_ISL_474030\|2020 | .................... | .................... | ..................... | .................... | .................... | .................... |
| D | 9340 hCoV-19/Wales/PHWC-163A75/2020\|EPI_ISL_474034\|2020 | .................... | .................... | ..................... | .................... | .................... | .................... |
| D | 9341 hCoV-19/Wales/PHWC-163A93/2020\|EPI_ISL_474036\|2020 | .................... | .................... | ..................... | .................... | .................... | .................... |
| D | 9342 hCoV-19/Wales/PHWC-163ADF/2020\|EPI_ISL_474040\|2020 | .................... | .................... | ..................... | .................... | .................... | .................... |
| D | 9343 hCoV-19/Wales/PHWC-163B18/2020\|EPI_ISL_474043\|2020 | .................... | .................... | ..................... | .................... | .................... | .................... |
| D | 9344 hCoV-19/Wales/PHWC-163B27/2020\|EPI_ISL_474044\|2020 | .................... | .................... | ..................... | .................... | .................... | .................... |
| D | 9345 hCoV-19/Wales/PHWC-163B54/2020\|EPI_ISL_474046\|2020 | .................... | .................... | ..................... | .................... | .................... | .................... |
| D | 9346 hCoV-19/Wales/PHWC-163B63/2020\|EPI_ISL_474047\|2020 | .................... | .................... | ..................... | .................... | .................... | .................... |
| D | 9347 hCoV-19/Wales/PHWC-163B72/2020\|EPI_ISL_474048\|2020 | .................... | .................... | ..................... | .................... | .................... | .................... |
| D | 9348 hCoV-19/Wales/PHWC-163B90/2020\|EPI_ISL_474050\|2020 | .................... | .................... | ..................... | .................... | .................... | .................... |
| D | 9349 hCoV-19/Wales/PHWC-163BBE/2020\|EPI_ISL_474052\|2020 | .................... | .................... | ..................... | .................... | .................... | .................... |
| D | 9350 hCoV-19/Wales/PHWC-163BEB/2020\|EPI_ISL_474055\|2020 | .................... | .................... | ..................... | .................... | .................... | .................... |
| D | 9351 hCoV-19/Wales/PHWC-163C06/2020\|EPI_ISL_474056\|2020 | .................... | .................... | ..................... | .................... | .................... | .................... |
| D | 9352 hCoV-19/Wales/PHWC-163D4F/2020\|EPI_ISL_474075\|2020 | .................... | .................... | ..................... | .................... | .................... | .................... |
| D | 9353 hCoV-19/Wales/PHWC-163D9A/2020\|EPI_ISL_474080\|2020 | .................... | .................... | ..................... | .................... | .................... | .................... |
| D | 9354 hCoV-19/Wales/PHWC-163DA9/2020\|EPI_ISL_474081\|2020 | .................... | .................... | ..................... | .................... | .................... | .................... |
| D | 9355 hCoV-19/Wales/PHWC-163DD6/2020\|EPI_ISL_474084\|2020 | .................... | .................... | ..................... | .................... | .................... | .................... |
| D | 9356 hCoV-19/Wales/PHWC-163DE5/2020\|EPI_ISL_474085\|2020 | .................... | .................... | ..................... | .................... | .................... | .................... |
| D | 9357 hCoV-19/Wales/PHWC-163E88/2020\|EPI_ISL_474095\|2020 | .................... | .................... | ..................... | .................... | .................... | .................... |
| D | 9358 hCoV-19/Wales/PHWC-163E97/2020\|EPI_ISL_474096\|2020 | .................... | .................... | ..................... | .................... | .................... | .................... |
| D | 9359 hCoV-19/Wales/PHWC-163EC4/2020\|EPI_ISL_474098\|2020 | .................... | .................... | ..................... | .................... | .................... | .................... |
| D | 9360 hCoV-19/Wales/PHWC-163ED3/2020\|EPI_ISL_474099\|2020 | .................... | .................... | ..................... | .................... | .................... | .................... |
| D | 9361 hCoV-19/England/PORT-2D0EB2/2020\|EPI_ISL_475300\|20 | .................... | .................... | ..................... | .................... | .................... | .................... |
| D | 9362 hCoV-19/England/PORT-2D0EEF/2020\|EPI_ISL_475302\|20 | .................... | .................... | ..................... | .................... | .................... | .................... |
| D | 9363 hCoV-19/England/PORT-2D0EFE/2020\|EPI_ISL_475303\|20 | .................... | .................... | ..................... | .................... | .................... | .................... |
| D | 9364 hCoV-19/England/PORT-2D0F0A/2020\|EPI_ISL_475304\|20 | .................... | .................... | ..................... | .................... | .................... | .................... |
| D | 9365 hCoV-19/England/PORT-2D0F28/2020\|EPI_ISL_475306\|20 | .................... | .................... | ..................... | .................... | .................... | .................... |
| D | 9366 hCoV-19/England/PORT-2D0F37/2020\|EPI_ISL_475307\|20 | .................... | .................... | ..................... | .................... | .................... | .................... |
| D | 9367 hCoV-19/England/PORT-2D0F64/2020\|EPI_ISL_475310\|20 | .................... | .................... | ..................... | .................... | .................... | .................... |
| D | 9368 hCoV-19/England/PORT-2D0F73/2020\|EPI_ISL_475311\|20 | .................... | .................... | ..................... | .................... | .................... | .................... |
| D | 9369 hCoV-19/England/PORT-2D0F82/2020\|EPI_ISL_475312\|20 | .................... | .................... | ..................... | .................... | .................... | .................... |
| D | 9370 hCoV-19/England/PORT-2D0FBF/2020\|EPI_ISL_475315\|20 | .................... | .................... | ..................... | .................... | .................... | .................... |
| D | 9371 hCoV-19/England/PORT-2D0FCE/2020\|EPI_ISL_475316\|20 | .................... | .................... | ..................... | .................... | .................... | .................... |
| D | 9372 hCoV-19/England/PORT-2D0FFB/2020\|EPI_ISL_475318\|20 | .................... | .................... | ..................... | .................... | .................... | .................... |
| D | 9373 hCoV-19/England/PORT-2D1024/2020\|EPI_ISL_475319\|20 | .................... | .................... | ..................... | .................... | .................... | .................... |
| D | 9374 hCoV-19/England/PORT-2D1042/2020\|EPI_ISL_475320\|20 | .................... | .................... | ..................... | .................... | .................... | .................... |
| D | 9375 hCoV-19/England/PORT-2D1060/2020\|EPI_ISL_475321\|20 | .................... | .................... | ..................... | .................... | .................... | .................... |
| D | 9376 hCoV-19/England/PORT-2D109D/2020\|EPI_ISL_475322\|20 | .................... | .................... | ..................... | .................... | .................... | .................... |
| D | 9377 hCoV-19/England/PORT-2D10AC/2020\|EPI_ISL_475323\|20 | .................... | .................... | ..................... | .................... | .................... | .................... |
| D | 9378 hCoV-19/England/PORT-2D10D9/2020\|EPI_ISL_475325\|20 | .................... | .................... | ..................... | .................... | .................... | .................... |
| D | 9379 hCoV-19/England/PORT-2D10E8/2020\|EPI_ISL_475326\|20 | .................... | .................... | ..................... | .................... | .................... | .................... |
| D | 9380 hCoV-19/England/PORT-2D10F7/2020\|EPI_ISL_475327\|20 | .................... | .................... | ..................... | .................... | .................... | .................... |
| D | 9381 hCoV-19/England/PORT-2D1103/2020\|EPI_ISL_475328\|20 | .................... | .................... | ..................... | .................... | .................... | .................... |
| D | 9382 hCoV-19/England/PORT-2D1112/2020\|EPI_ISL_475329\|20 | .................... | .................... | ..................... | .................... | .................... | .................... |
| D | 9383 hCoV-19/England/PORT-2D1121/2020\|EPI_ISL_475330\|20 | .................... | .................... | ..................... | .................... | .................... | .................... |
| D | 9384 hCoV-19/England/PORT-2D1130/2020\|EPI_ISL_475331\|20 | .................... | .................... | ..................... | .................... | .................... | .................... |
| D | 9385 hCoV-19/England/PORT-2D116D/2020\|EPI_ISL_475333\|20 | .................... | .................... | ..................... | .................... | .................... | .................... |
| D | 9386 hCoV-19/England/PORT-2D117C/2020\|EPI_ISL_475334\|20 | .................... | .................... | ..................... | .................... | .................... | .................... |
| D | 9387 hCoV-19/England/PORT-2D118B/2020\|EPI_ISL_475335\|20 | .................... | .................... | ..................... | .................... | .................... | .................... |
| D | 9388 hCoV-19/England/PORT-2D11A9/2020\|EPI_ISL_475337\|20 | .................... | .................... | ..................... | .................... | .................... | .................... |
| D | 9389 hCoV-19/England/PORT-2D1200/2020\|EPI_ISL_475339\|20 | .................... | .................... | ..................... | .................... | .................... | .................... |
| D | 9390 hCoV-19/England/PORT-2D12A6/2020\|EPI_ISL_475341\|20 | .................... | .................... | ..................... | .................... | .................... | .................... |
| D | 9391 hCoV-19/England/SHEF-C09D4/2020\|EPI_ISL_475343\|202 | .................... | .................... | ..................... | .................... | .................... | .................... |
| D | 9392 hCoV-19/England/SHEF-C0A1D/2020\|EPI_ISL_475344\|202 | .................... | .................... | ..................... | .................... | .................... | .................... |
| D | 9393 hCoV-19/England/SHEF-C0BCF/2020\|EPI_ISL_475347\|202 | .................... | .................... | ..................... | .................... | .................... | .................... |
| D | 9394 hCoV-19/England/SHEF-C0D50/2020\|EPI_ISL_475348\|202 | .................... | .................... | ..................... | .................... | .................... | .................... |
| D | 9395 hCoV-19/England/SHEF-C1EF2/2020\|EPI_ISL_475356\|202 | .................... | .................... | ..................... | .................... | .................... | .................... |
| D | 9396 hCoV-19/England/SHEF-C232F/2020\|EPI_ISL_475360\|202 | .................... | .................... | ..................... | .................... | .................... | .................... |
| D | 9397 hCoV-19/England/SHEF-C2969/2020\|EPI_ISL_475367\|202 | .................... | .................... | ..................... | .................... | .................... | .................... |
| D | 9398 hCoV-19/England/SHEF-C335B/2020\|EPI_ISL_475368\|202 | .................... | .................... | ..................... | .................... | .................... | .................... |
| D | 9399 hCoV-19/England/SHEF-C3986/2020\|EPI_ISL_475370\|202 | .................... | .................... | ..................... | .................... | .................... | .................... |

| | | | | | | | | |
|---|---|---|---|---|---|---|---|---|
| D | 9400 hCoV-19/England/SHEF-C3A0B/2020\|EPI_ISL_475371\|202 | .................... | .................... | .................... | .................... | .................... | .................... | |
| D | 9401 hCoV-19/England/SHEF-C3B08/2020\|EPI_ISL_475372\|202 | .................... | .................... | .................... | .................... | .................... | .................... | |
| D | 9402 hCoV-19/England/SHEF-C3F1B/2020\|EPI_ISL_475375\|202 | .................... | .................... | .................... | .................... | .................... | .................... | |
| D | 9403 hCoV-19/England/SHEF-C4C5F/2020\|EPI_ISL_475378\|202 | .................... | .................... | .................... | .................... | .................... | .................... | |
| D | 9404 hCoV-19/England/SHEF-C4C8C/2020\|EPI_ISL_475379\|202 | .................... | .................... | .................... | .................... | .................... | .................... | |
| D | 9405 hCoV-19/England/SHEF-C58A5/2020\|EPI_ISL_475384\|202 | .................... | .................... | .................... | .................... | .................... | .................... | |
| D | 9406 hCoV-19/Scotland/CVR2371/2020\|EPI_ISL_477920\|2020- | .................... | .................... | .................... | .................... | .................... | .................... | |
| D | 9407 hCoV-19/Scotland/CVR2382/2020\|EPI_ISL_477922\|2020- | .................... | .................... | .................... | .................... | .................... | .................... | |
| D | 9408 hCoV-19/Scotland/CVR2384/2020\|EPI_ISL_477924\|2020- | .................... | .................... | .................... | .................... | .................... | .................... | |
| D | 9409 hCoV-19/Scotland/CVR2385/2020\|EPI_ISL_477925\|2020- | .................... | .................... | .................... | .................... | .................... | .................... | |
| D | 9410 hCoV-19/Scotland/CVR2390/2020\|EPI_ISL_477926\|2020- | .................... | .................... | .................... | .................... | .................... | .................... | |
| D | 9411 hCoV-19/Scotland/CVR2395/2020\|EPI_ISL_477928\|2020- | .................... | .................... | .................... | .................... | .................... | .................... | |
| D | 9412 hCoV-19/Scotland/CVR2396/2020\|EPI_ISL_477929\|2020- | .................... | .................... | .................... | .................... | .................... | .................... | |
| D | 9413 hCoV-19/Scotland/CVR2397/2020\|EPI_ISL_477930\|2020- | .................... | .................... | .................... | .................... | .................... | .................... | |
| D | 9414 hCoV-19/Scotland/CVR2405/2020\|EPI_ISL_477932\|2020- | .................... | ..............C..... | .................... | .................... | .................... | .................... | |
| D | 9415 hCoV-19/Scotland/CVR2416/2020\|EPI_ISL_477933\|2020- | .................... | .................... | .................... | .................... | .................... | .................... | |
| D | 9416 hCoV-19/Scotland/CVR2417/2020\|EPI_ISL_477934\|2020- | .................... | .A.................. | .................... | .................... | .................... | .................... | |
| D | 9417 hCoV-19/Scotland/CVR2420/2020\|EPI_ISL_477935\|2020- | .................... | .................... | .................... | .................... | .................... | .................... | |
| D | 9418 hCoV-19/Scotland/CVR2421/2020\|EPI_ISL_477936\|2020- | .................... | .................... | .................... | .................... | .................... | .................... | |
| D | 9419 hCoV-19/Scotland/CVR2429/2020\|EPI_ISL_477938\|2020- | .................... | ...................A | .................... | .................... | .................... | .................... | 1 |
| D | 9420 hCoV-19/Scotland/CVR2441/2020\|EPI_ISL_477939\|2020- | .................... | .................... | .................... | .................... | .................... | .................... | |
| D | 9421 hCoV-19/Scotland/CVR2442/2020\|EPI_ISL_477940\|2020- | .................... | .................... | .................... | .................... | .................... | .................... | |
| D | 9422 hCoV-19/Scotland/CVR2444/2020\|EPI_ISL_477941\|2020- | .................... | .A.................. | .................... | .................... | .................... | .................... | |
| D | 9423 hCoV-19/Scotland/CVR2445/2020\|EPI_ISL_477942\|2020- | .................... | .................... | .................... | .................... | .................... | .................... | |
| D | 9424 hCoV-19/Scotland/CVR2448/2020\|EPI_ISL_477943\|2020- | .................... | .................... | .................... | .................... | .................... | .................... | |
| D | 9425 hCoV-19/Scotland/CVR2453/2020\|EPI_ISL_477944\|2020- | .................... | .................... | .................... | .................... | .................... | .................... | |
| D | 9426 hCoV-19/Scotland/CVR2458/2020\|EPI_ISL_477946\|2020- | .................... | .................... | .................... | .................... | .................... | .................... | |
| D | 9427 hCoV-19/Scotland/CVR2460/2020\|EPI_ISL_477947\|2020- | .................... | .................... | .................... | .................... | .................... | .................... | |
| D | 9428 hCoV-19/Scotland/CVR2469/2020\|EPI_ISL_477949\|2020- | .................... | .................... | .................... | .................... | .................... | .................... | |
| D | 9429 hCoV-19/Scotland/CVR2480/2020\|EPI_ISL_477951\|2020- | .................... | ...................A | .................... | .................... | .................... | .................... | 1 |
| D | 9430 hCoV-19/Scotland/CVR2484/2020\|EPI_ISL_477952\|2020- | .................... | .................... | .................... | .................... | .................... | .................... | |
| D | 9431 hCoV-19/Scotland/CVR2488/2020\|EPI_ISL_477953\|2020- | .................... | .................... | .................... | .................... | .................... | .................... | |
| D | 9432 hCoV-19/Scotland/CVR2490/2020\|EPI_ISL_477954\|2020- | .................... | .................... | .................... | .................... | .................... | .................... | |
| D | 9433 hCoV-19/Scotland/CVR2494/2020\|EPI_ISL_477955\|2020- | .................... | .................... | .................... | .................... | .................... | .................... | |
| D | 9434 hCoV-19/Scotland/CVR2495/2020\|EPI_ISL_477956\|2020- | .................... | .................... | .................... | .................... | .................... | .................... | |
| D | 9435 hCoV-19/Scotland/CVR2498/2020\|EPI_ISL_477957\|2020- | .................... | .................... | .................... | .................... | .................... | .................... | |
| D | 9436 hCoV-19/Scotland/CVR2503/2020\|EPI_ISL_477958\|2020- | .................... | .................... | .................... | .................... | .................... | .................... | |
| D | 9437 hCoV-19/Scotland/CVR2517/2020\|EPI_ISL_477960\|2020- | .................... | .................... | .................... | .................... | .................... | .................... | |
| D | 9438 hCoV-19/Scotland/CVR2520/2020\|EPI_ISL_477961\|2020- | .................... | ...................A | .................... | .................... | .................... | .................... | 1 |
| D | 9439 hCoV-19/Scotland/CVR2523/2020\|EPI_ISL_477962\|2020- | .................... | ...................A | .................... | .................... | .................... | .................... | 1 |
| D | 9440 hCoV-19/Scotland/CVR2534/2020\|EPI_ISL_477964\|2020- | .................... | .A.................. | .................... | .................... | .................... | .................... | |
| D | 9441 hCoV-19/Scotland/CVR2538/2020\|EPI_ISL_477965\|2020- | .................... | ...................A | .................... | .................... | .................... | .................... | 1 |
| D | 9442 hCoV-19/Scotland/CVR2554/2020\|EPI_ISL_477967\|2020- | .................... | .A.................. | .................... | .................... | .................... | .................... | |
| D | 9443 hCoV-19/Scotland/CVR2561/2020\|EPI_ISL_477969\|2020- | .................... | .................... | .................... | .................... | .................... | .................... | |
| D | 9444 hCoV-19/Scotland/CVR2562/2020\|EPI_ISL_477970\|2020- | .................... | .................... | .................... | .................... | .................... | .................... | |
| D | 9445 hCoV-19/Scotland/CVR2563/2020\|EPI_ISL_477971\|2020- | .................... | ...................A | .................... | .................... | .................... | .................... | 1 |
| D | 9446 hCoV-19/Scotland/CVR2564/2020\|EPI_ISL_477972\|2020- | .................... | ..............C..... | .................... | .................... | .................... | .................... | |
| D | 9447 hCoV-19/Scotland/CVR2566/2020\|EPI_ISL_477973\|2020- | .................... | .................... | .................... | .................... | .................... | .................... | |
| D | 9448 hCoV-19/Scotland/CVR2568/2020\|EPI_ISL_477974\|2020- | .................... | ...................A | .................... | .................... | .................... | .................... | 1 |
| D | 9449 hCoV-19/Scotland/CVR2571/2020\|EPI_ISL_477975\|2020- | .................... | .................... | .................... | .................... | .................... | .................... | |
| D | 9450 hCoV-19/Scotland/CVR2573/2020\|EPI_ISL_477976\|2020- | .................... | .................... | .................... | .................... | .................... | .................... | |
| D | 9451 hCoV-19/Scotland/CVR2574/2020\|EPI_ISL_477977\|2020- | .................... | .................... | .................... | .................... | .................... | .................... | |
| D | 9452 hCoV-19/Scotland/CVR2575/2020\|EPI_ISL_477978\|2020- | .................... | ...................A | .................... | .................... | .................... | .................... | 1 |
| D | 9453 hCoV-19/Scotland/CVR2578/2020\|EPI_ISL_477979\|2020- | .................... | .................... | .................... | .................... | .................... | .................... | |
| D | 9454 hCoV-19/Scotland/CVR2592/2020\|EPI_ISL_477980\|2020- | .................... | ..............C..... | .................... | .................... | .................... | .................... | |
| D | 9455 hCoV-19/Scotland/CVR2593/2020\|EPI_ISL_477981\|2020- | .................... | .................... | .................... | .................... | .................... | .................... | |
| D | 9456 hCoV-19/Scotland/CVR2597/2020\|EPI_ISL_477983\|2020- | .................... | .................... | .................... | .................... | .................... | .................... | |
| D | 9457 hCoV-19/Scotland/CVR2600/2020\|EPI_ISL_477984\|2020- | .................... | .................... | .................... | .................... | .................... | .................... | |
| D | 9458 hCoV-19/Scotland/CVR2602/2020\|EPI_ISL_477985\|2020- | .................... | ...................A | .................... | .................... | .................... | .................... | 1 |
| D | 9459 hCoV-19/Scotland/CVR2604/2020\|EPI_ISL_477986\|2020- | .................... | .................... | .................... | .................... | .................... | .................... | |
| D | 9460 hCoV-19/Scotland/CVR2610/2020\|EPI_ISL_477987\|2020- | .................... | .................... | .................... | .................... | .................... | .................... | |
| D | 9461 hCoV-19/Scotland/CVR2616/2020\|EPI_ISL_477988\|2020- | .................... | ...................A | .................... | .................... | .................... | .................... | 1 |
| D | 9462 hCoV-19/Scotland/CVR2619/2020\|EPI_ISL_477989\|2020- | .................... | .................... | .................... | .................... | .................... | .................... | |
| D | 9463 hCoV-19/Scotland/CVR2620/2020\|EPI_ISL_477990\|2020- | .................... | .................... | .................... | .................... | .................... | .................... | |
| D | 9464 hCoV-19/Scotland/CVR2624/2020\|EPI_ISL_477991\|2020- | .................... | .................... | .................... | .................... | .................... | ...............A.... | |
| D | 9465 hCoV-19/Scotland/CVR2627/2020\|EPI_ISL_477992\|2020- | .................... | .................... | .................... | .................... | .................... | .................... | |
| D | 9466 hCoV-19/Scotland/CVR2628/2020\|EPI_ISL_477993\|2020- | .................... | ...................A | .................... | .................... | .................... | .................... | 1 |
| D | 9467 hCoV-19/Scotland/CVR2629/2020\|EPI_ISL_477994\|2020- | .................... | .A.................. | .................... | .................... | .................... | .................... | |
| D | 9468 hCoV-19/Scotland/CVR2631/2020\|EPI_ISL_477995\|2020- | .................... | .................... | .................... | .................... | .................... | .................... | |
| D | 9469 hCoV-19/Scotland/CVR2632/2020\|EPI_ISL_477996\|2020- | .................... | .................... | .................... | .................... | .................... | .................... | |
| D | 9470 hCoV-19/Scotland/CVR2635/2020\|EPI_ISL_477997\|2020- | .................... | .................... | .................... | .................... | .................... | .................... | |
| D | 9471 hCoV-19/Scotland/CVR2637/2020\|EPI_ISL_477998\|2020- | .................... | .................... | .................... | .................... | .................... | .................... | |
| D | 9472 hCoV-19/Scotland/CVR2638/2020\|EPI_ISL_477999\|2020- | .................... | .................... | .................... | .................... | .................... | .................... | |
| D | 9473 hCoV-19/England/20142005504/2020\|EPI_ISL_465904\|20 | .................... | .................... | .................... | .................... | .................... | .................... | |
| D | 9474 hCoV-19/England/20142006104/2020\|EPI_ISL_465906\|20 | .................... | .................... | .................... | .................... | .................... | .................... | |
| D | 9475 hCoV-19/England/20142010304/2020\|EPI_ISL_465914\|20 | .................... | .................... | .................... | .................... | .................... | .................... | |
| D | 9476 hCoV-19/England/20142014704/2020\|EPI_ISL_465920\|20 | .................... | .................... | .................... | .................... | .................... | .................... | |
| D | 9477 hCoV-19/England/20142014904/2020\|EPI_ISL_465921\|20 | .................... | .................... | .................... | .................... | .................... | .................... | |

| | | | | | | | | |
|---|---|---|---|---|---|---|---|---|
| D | 9478 | hCoV-19/England/20142015404/2020\|EPI_ISL_465922\|20 | ···················· | ····A··············· | ····················· | ···················· | ···················· | ···················· |
| D | 9479 | hCoV-19/England/20142016604/2020\|EPI_ISL_465923\|20 | ···················· | ···················· | ····················· | ···················· | ···················· | ···················· |
| D | 9480 | hCoV-19/England/20142018404/2020\|EPI_ISL_465926\|20 | ···················· | ···················· | ····················· | ···················· | ···················· | ···················· |
| D | 9481 | hCoV-19/England/20142022204/2020\|EPI_ISL_465932\|20 | ···················· | ···················· | ····················· | ···················· | ···················· | ···················· |
| D | 9482 | hCoV-19/England/20142024504/2020\|EPI_ISL_465935\|20 | ···················· | ···················· | ····················· | ···················· | ···················· | ···················· |
| D | 9483 | hCoV-19/England/20142025204/2020\|EPI_ISL_465936\|20 | ···················· | ···················· | ····················· | ···················· | ···················· | ···················· |
| D | 9484 | hCoV-19/England/20142025404/2020\|EPI_ISL_465937\|20 | ···················· | ···················· | ····················· | ···················· | ···················· | ···················· |
| D | 9485 | hCoV-19/England/20142026704/2020\|EPI_ISL_465939\|20 | ···················· | ···················· | ····················· | ···················· | ···················· | ···················· |
| D | 9486 | hCoV-19/England/20144021904/2020\|EPI_ISL_465989\|20 | ···················· | ···················· | ····················· | ···················· | ···················· | ···················· |
| D | 9487 | hCoV-19/USA/OR-OHSU-0093/2020\|EPI_ISL_468103\|2020- | ···················· | ···················· | ····················· | ···················· | ···················· | ···················· |
| D | 9488 | hCoV-19/USA/OR-OHSU-0135/2020\|EPI_ISL_468109\|2020- | ···················· | ···················· | ····················· | ···················· | ···················· | ···················· |
| D | 9489 | hCoV-19/USA/OR-OHSU-0140/2020\|EPI_ISL_468110\|2020- | ···················· | ···················· | ····················· | ···················· | ···················· | ···················· |
| D | 9490 | hCoV-19/USA/OR-OHSU-0150/2020\|EPI_ISL_468112\|2020- | ···················· | ···················· | ····················· | ···················· | ···················· | ···················· |
| D | 9491 | hCoV-19/USA/OR-OHSU-0151/2020\|EPI_ISL_468113\|2020- | ···················· | ···················· | ····················· | ···················· | ···················· | ···················· |
| D | 9492 | hCoV-19/USA/OR-OHSU-0173/2020\|EPI_ISL_468117\|2020- | ···················· | ···················· | ····················· | ···················· | ···················· | ···················· |
| D | 9493 | hCoV-19/USA/OR-OHSU-0177/2020\|EPI_ISL_468120\|2020- | ···················· | ···················· | ····················· | ···················· | ···················· | ···················· |
| D | 9494 | hCoV-19/USA/OR-OHSU-0181/2020\|EPI_ISL_468123\|2020- | ···················· | ···················· | ····················· | ···················· | ···················· | ···················· |
| D | 9495 | hCoV-19/USA/OR-OHSU-0183/2020\|EPI_ISL_468125\|2020- | ···················· | ···················· | ····················· | ···················· | ···················· | ···················· |
| D | 9496 | hCoV-19/USA/OR-OHSU-0184/2020\|EPI_ISL_468126\|2020- | ···················· | ···················· | ····················· | ···················· | ···················· | ···················· |
| D | 9497 | hCoV-19/Romania/Bucuresti-4105/2020\|EPI_ISL_468134 | ···················· | ···················· | ····················· | ···················· | ···················· | ···················· |
| D | 9498 | hCoV-19/Romania/Bucuresti-4284/2020\|EPI_ISL_468135 | ···················· | ···················· | ····················· | ···················· | ···················· | ···················· |
| D | 9499 | hCoV-19/Romania/Bucuresti-4322/2020\|EPI_ISL_468136 | ···················· | ···················· | ····················· | ···················· | ···················· | ···················· |
| D | 9500 | hCoV-19/Romania/Suceava-4682/2020\|EPI_ISL_468137\|2 | ···················· | ···················· | ····················· | ···················· | ···················· | ···················· |
| D | 9501 | hCoV-19/Romania/Suceava-5709/2020\|EPI_ISL_468138\|2 | ···················· | ···················· | ····················· | ···················· | ···················· | ···················· |
| D | 9502 | hCoV-19/Romania/Suceava-5710/2020\|EPI_ISL_468139\|2 | ···················· | ···················· | ····················· | ···················· | ···················· | ···················· |
| D | 9503 | hCoV-19/Romania/Suceava-5722/2020\|EPI_ISL_468140\|2 | ···················· | ···················· | ····················· | ···················· | ···················· | ···················· |
| D | 9504 | hCoV-19/Romania/Suceava-5806/2020\|EPI_ISL_468141\|2 | ···················· | ···················· | ····················· | ···················· | ···················· | ···················· |
| D | 9505 | hCoV-19/Romania/Suceava-5893/2020\|EPI_ISL_468143\|2 | ···················· | ···················· | ····················· | ···················· | ···················· | ···················· |
| D | 9506 | hCoV-19/Romania/Suceava-5894/2020\|EPI_ISL_468144\|2 | ···················· | ···················· | ····················· | ···················· | ···················· | ···················· |
| D | 9507 | hCoV-19/Romania/Bucuresti-6451/2020\|EPI_ISL_468145 | ···················· | ···················· | ····················· | ···················· | ···················· | ···················· |
| D | 9508 | hCoV-19/Romania/Suceava-11085/2020\|EPI_ISL_468146\| | ···················· | ···················· | ····················· | ···················· | ···················· | ···················· |
| D | 9509 | hCoV-19/Romania/Bucuresti-12292/2020\|EPI_ISL_46814 | ···················· | ···················· | ····················· | ···················· | ···················· | ···················· |
| D | 9510 | hCoV-19/Romania/Bucuresti-12296/2020\|EPI_ISL_46814 | ···················· | ···················· | ····················· | ···················· | ···················· | ···················· |
| D | 9511 | hCoV-19/Romania/Bucuresti-12546/2020\|EPI_ISL_46814 | ···················· | ···················· | ····················· | ···················· | ···················· | ···················· |
| D | 9512 | hCoV-19/Romania/Bucuresti-12547/2020\|EPI_ISL_46815 | ···················· | ···················· | ····················· | ···················· | ···················· | ···················· |
| D | 9513 | hCoV-19/Romania/Bucuresti-14545/2020\|EPI_ISL_46815 | ···················· | ···················· | ····················· | ···················· | ···················· | ···················· |
| D | 9514 | hCoV-19/Romania/Bucuresti-14658/2020\|EPI_ISL_46815 | ···················· | ···················· | ····················· | ···················· | ···················· | ···················· |
| D | 9515 | hCoV-19/Romania/Bucuresti-15503/2020\|EPI_ISL_46815 | ···················· | ···················· | ····················· | ···················· | ···················· | ···················· |
| D | 9516 | hCoV-19/Romania/Mioveni-24095/2020\|EPI_ISL_468156\| | ···················· | ···················· | ····················· | ···················· | ···················· | ···················· |
| D | 9517 | hCoV-19/Romania/Suceava-31646/2020\|EPI_ISL_468158\| | ···················· | ···················· | ····················· | ···················· | ···················· | ···················· |
| D | 9518 | hCoV-19/Pakistan/NIH-44905/2020\|EPI_ISL_468159\|202 | ···················· | ···················· | ····················· | ···················· | ···················· | ···················· |
| D | 9519 | hCoV-19/Pakistan/NIH-45143/2020\|EPI_ISL_468160\|202 | ···················· | ···················· | ····················· | ···················· | ···················· | ···················· |
| D | 9520 | hCoV-19/Pakistan/NIH-45090/2020\|EPI_ISL_468161\|202 | ···················· | ···················· | ····················· | ···················· | ···················· | ···················· |
| D | 9521 | hCoV-19/Pakistan/NIH-45579/2020\|EPI_ISL_468162\|202 | ···················· | ···················· | ····················· | ···················· | ···················· | ···················· |
| D | 9522 | hCoV-19/Pakistan/NIH-HAS001/2020\|EPI_ISL_468163\|20 | ···················· | ···················· | ····················· | ···················· | ···················· | ···················· |
| D | 9523 | hCoV-19/England/BRIS-12BA79/2020\|EPI_ISL_469400\|20 | ···················· | ···················· | ····················· | ···················· | ···················· | ···················· |
| D | 9524 | hCoV-19/England/BRIS-12BB94/2020\|EPI_ISL_469401\|20 | ···················· | ···················· | ····················· | ···················· | ···················· | ···················· |
| D | 9525 | hCoV-19/England/BRIS-12BE5F/2020\|EPI_ISL_469402\|20 | ···················· | ···················· | ····················· | ···················· | ···················· | ···················· |
| D | 9526 | hCoV-19/England/BRIS-12C155/2020\|EPI_ISL_469403\|20 | ···················· | ···················· | ····················· | ···················· | ···················· | ···················· |
| D | 9527 | hCoV-19/England/BRIS-12BA88/2020\|EPI_ISL_469404\|20 | ···················· | ···················· | ····················· | ···················· | ···················· | ···················· |
| D | 9528 | hCoV-19/England/BRIS-12C39B/2020\|EPI_ISL_469405\|20 | ···················· | ···················· | ····················· | ···················· | ···················· | ···················· |
| D | 9529 | hCoV-19/England/BRIS-12B8E8/2020\|EPI_ISL_469406\|20 | ···················· | ···················· | ····················· | ···················· | ···················· | ···················· |
| D | 9530 | hCoV-19/England/BRIS-12C137/2020\|EPI_ISL_469407\|20 | ···················· | ···················· | ····················· | ···················· | ···················· | ···················· |
| D | 9531 | hCoV-19/England/BRIS-12BC82/2020\|EPI_ISL_469408\|20 | ···················· | ···················· | ····················· | ···················· | ···················· | ···················· |
| D | 9532 | hCoV-19/England/BRIS-12BF89/2020\|EPI_ISL_469409\|20 | ···················· | ···················· | ····················· | ···················· | ···················· | ···················· |
| D | 9533 | hCoV-19/England/BRIS-12C28F/2020\|EPI_ISL_469410\|20 | ···················· | ·A·················· | ····················· | ···················· | ···················· | ···················· |
| D | 9534 | hCoV-19/England/BRIS-12C270/2020\|EPI_ISL_469412\|20 | ···················· | ·A·················· | ····················· | ···················· | ···················· | ···················· |
| D | 9535 | hCoV-19/England/BRIS-12B9C7/2020\|EPI_ISL_469413\|20 | ···················· | ···················· | ····················· | ···················· | ···················· | ···················· |
| D | 9536 | hCoV-19/England/BRIS-12C03A/2020\|EPI_ISL_469414\|20 | ···················· | ···················· | ····················· | ···················· | ···················· | ···················· |
| D | 9537 | hCoV-19/England/BRIS-12BC28/2020\|EPI_ISL_469415\|20 | ···················· | ···················· | ····················· | ···················· | ···················· | ···················· |
| D | 9538 | hCoV-19/England/BRIS-12B97C/2020\|EPI_ISL_469416\|20 | ···················· | ···················· | ····················· | ···················· | ···················· | ···················· |
| D | 9539 | hCoV-19/England/BRIS-12C38C/2020\|EPI_ISL_469417\|20 | ···················· | ···················· | ····················· | ···················· | ···················· | ···················· |
| D | 9540 | hCoV-19/England/BRIS-12BAE2/2020\|EPI_ISL_469418\|20 | ···················· | ···················· | ····················· | ···················· | ···················· | ···················· |
| D | 9541 | hCoV-19/England/BRIS-12B9A9/2020\|EPI_ISL_469419\|20 | ···················· | ···················· | ····················· | ···················· | ···················· | ···················· |
| D | 9542 | hCoV-19/England/BRIS-12C43E/2020\|EPI_ISL_469420\|20 | ···················· | ···················· | ····················· | ···················· | ···················· | ···················· |
| D | 9543 | hCoV-19/England/BRIS-12C234/2020\|EPI_ISL_469421\|20 | ···················· | ·A·················· | ····················· | ···················· | ···················· | ···················· |
| D | 9544 | hCoV-19/England/BRIS-12C1BF/2020\|EPI_ISL_469422\|20 | ···················· | ···················· | ····················· | ···················· | ···················· | ···················· |
| D | 9545 | hCoV-19/England/BRIS-12C128/2020\|EPI_ISL_469423\|20 | ···················· | ···················· | ····················· | ···················· | ···················· | ···················· |
| D | 9546 | hCoV-19/England/BRIS-12BD9E/2020\|EPI_ISL_469424\|20 | ···················· | ···················· | ····················· | ···················· | ···················· | ···················· |
| D | 9547 | hCoV-19/England/BRIS-12C076/2020\|EPI_ISL_469430\|20 | ···················· | ···················· | ····················· | ···················· | ···················· | ···················· |
| D | 9548 | hCoV-19/England/BRIS-12BAD3/2020\|EPI_ISL_469431\|20 | ···················· | ···················· | ····················· | ···················· | ···················· | ···················· |
| D | 9549 | hCoV-19/England/BRIS-12B88E/2020\|EPI_ISL_469432\|20 | ···················· | ···················· | ····················· | ···················· | ···················· | ···················· |
| D | 9550 | hCoV-19/England/BRIS-12C00D/2020\|EPI_ISL_469436\|20 | ···················· | ···················· | ····················· | ···················· | ···················· | ···················· |
| D | 9551 | hCoV-19/England/BRIS-12C243/2020\|EPI_ISL_469437\|20 | ···················· | ·A·················· | ····················· | ···················· | ···················· | ···················· |
| D | 9552 | hCoV-19/England/BRIS-12B9D6/2020\|EPI_ISL_469438\|20 | ···················· | ···················· | ····················· | ···················· | ···················· | ···················· |
| D | 9553 | hCoV-19/England/BRIS-12BF2F/2020\|EPI_ISL_469439\|20 | ···················· | ···················· | ····················· | ···················· | ···················· | ···················· |
| D | 9554 | hCoV-19/England/BRIS-12BC64/2020\|EPI_ISL_469440\|20 | ···················· | ···················· | ····················· | ···················· | ···················· | ···················· |
| D | 9555 | hCoV-19/England/BRIS-12BB3A/2020\|EPI_ISL_469442\|20 | ···················· | ···················· | ····················· | ···················· | ···················· | ···················· |

D 9556 hCoV-19/England/BRIS-12BEF5/2020|EPI_ISL_469443|20 ···················· ···················· ···················· ···················· ···················· ····················
D 9557 hCoV-19/England/BRIS-12BC46/2020|EPI_ISL_469444|20 ···················· ···················· ···················· ···················· ···················· ····················
D 9558 hCoV-19/England/BRIS-12C2BC/2020|EPI_ISL_469445|20 ···················· ·A·················· ···················· ···················· ···················· ····················
D 9559 hCoV-19/England/BRIS-12BC0A/2020|EPI_ISL_469446|20 ···················· ···················· ···················· ···················· ···················· ····················
D 9560 hCoV-19/England/BRIS-12BA00/2020|EPI_ISL_469448|20 ···················· ···················· ···················· ···················· ···················· ····················
D 9561 hCoV-19/England/BRIS-12BCEC/2020|EPI_ISL_469449|20 ···················· ···················· ···················· ···················· ···················· ····················
D 9562 hCoV-19/England/BRIS-12C37D/2020|EPI_ISL_469451|20 ···················· ···················· ···················· ···················· ···················· ····················
D 9563 hCoV-19/England/BRIS-12B98B/2020|EPI_ISL_469452|20 ···················· ···················· ···················· ···················· ···················· ····················
D 9564 hCoV-19/England/BRIS-12C085/2020|EPI_ISL_469453|20 ···················· ···················· ···················· ···················· ···················· ····················
D 9565 hCoV-19/England/BRIS-12C058/2020|EPI_ISL_469454|20 ···················· ···················· ···················· ···················· ···················· ····················
D 9566 hCoV-19/England/BRIS-12BBEF/2020|EPI_ISL_469455|20 ···················· ···················· ···················· ···················· ···················· ····················
D 9567 hCoV-19/England/BRIS-12C3F5/2020|EPI_ISL_469456|20 ···················· ···················· ···················· ···················· ···················· ····················
D 9568 hCoV-19/England/BRIS-12BB85/2020|EPI_ISL_469457|20 ···················· ···················· ···················· ···················· ···················· ····················
D 9569 hCoV-19/England/BRIS-12BF10/2020|EPI_ISL_469458|20 ···················· ···················· ···················· ···················· ···················· ····················
D 9570 hCoV-19/England/BRIS-12C29E/2020|EPI_ISL_469459|20 ···················· ·A·················· ···················· ···················· ···················· ····················
D 9571 hCoV-19/England/BRIS-12C01C/2020|EPI_ISL_469462|20 ···················· ···················· ···················· ···················· ···················· ····················
D 9572 hCoV-19/England/BRIS-12BDCB/2020|EPI_ISL_469463|20 ···················· ···················· ···················· ···················· ···················· ····················
D 9573 hCoV-19/England/BRIS-12C638/2020|EPI_ISL_469464|20 ···················· ···················· ···················· ···················· ···················· ····················
D 9574 hCoV-19/England/BRIS-12C6CF/2020|EPI_ISL_469465|20 ···················· ···················· ···················· ···················· ···················· ····················
D 9575 hCoV-19/England/BRIS-12C568/2020|EPI_ISL_469466|20 ···················· ···················· ···················· ···················· ···················· ····················
D 9576 hCoV-19/England/BRIS-12C726/2020|EPI_ISL_469468|20 ···················· ···················· ···················· ···················· ···················· ····················
D 9577 hCoV-19/England/BRIS-12C586/2020|EPI_ISL_469470|20 ···················· ···················· ···················· ···················· ···················· ····················
D 9578 hCoV-19/England/BRIS-12C5E0/2020|EPI_ISL_469471|20 ···················· ···················· ···················· ···················· ···················· ····················
D 9579 hCoV-19/England/BRIS-12C87E/2020|EPI_ISL_469472|20 ···················· ···················· ···················· ···················· ···················· ····················
D 9580 hCoV-19/England/BRIS-12C7F9/2020|EPI_ISL_469474|20 ···················· ···················· ···················· ···················· ···················· ····················
D 9581 hCoV-19/England/BRIS-12C577/2020|EPI_ISL_469475|20 ···················· ···················· ···················· ···················· ···················· ····················
D 9582 hCoV-19/England/BRIS-12C4D4/2020|EPI_ISL_469476|20 ···················· ···················· ···················· ···················· ···················· ····················
D 9583 hCoV-19/England/BRIS-12C692/2020|EPI_ISL_469477|20 ···················· ···················· ···················· ···················· ···················· ····················
D 9584 hCoV-19/England/BRIS-12C6B0/2020|EPI_ISL_469478|20 ···················· ···················· ···················· ···················· ···················· ····················
D 9585 hCoV-19/England/BRIS-12C4C5/2020|EPI_ISL_469479|20 ···················· ···················· ···················· ···················· ···················· ····················
D 9586 hCoV-19/England/BRIS-12C5FF/2020|EPI_ISL_469480|20 ···················· ···················· ···················· ···················· ···················· ····················
D 9587 hCoV-19/England/BRIS-12C708/2020|EPI_ISL_469481|20 ···················· ···················· ···················· ···················· ···················· ····················
D 9588 hCoV-19/England/BRIS-12C647/2020|EPI_ISL_469483|20 ···················· ···················· ···················· ···················· ···················· ····················
D 9589 hCoV-19/England/BRIS-12C656/2020|EPI_ISL_469484|20 ···················· ···················· ···················· ···················· ···················· ····················
D 9590 hCoV-19/England/BRIS-12C54A/2020|EPI_ISL_469485|20 ···················· ···················· ···················· ···················· ···················· ····················
D 9591 hCoV-19/England/BRIS-12C52C/2020|EPI_ISL_469486|20 ···················· ···················· ···················· ···················· ···················· ····················
D 9592 hCoV-19/England/BRIS-12C60B/2020|EPI_ISL_469488|20 ···················· ···················· ···················· ···················· ···················· ····················
D 9593 hCoV-19/England/BRIS-12C6ED/2020|EPI_ISL_469489|20 ···················· ···················· ···················· ···················· ···················· ····················
D 9594 hCoV-19/England/BRIS-12C753/2020|EPI_ISL_469490|20 ···················· ···················· ···················· ···················· ···················· ····················
D 9595 hCoV-19/England/BRIS-12C7EA/2020|EPI_ISL_469492|20 ···················· ···················· ···················· ···················· ···················· ····················
D 9596 hCoV-19/England/BRIS-12C498/2020|EPI_ISL_469493|20 ···················· ···················· ···················· ···················· ···················· ····················
D 9597 hCoV-19/England/BRIS-12C51D/2020|EPI_ISL_469494|20 ···················· ···················· ···················· ···················· ···················· ····················
D 9598 hCoV-19/England/BRIS-12C8BA/2020|EPI_ISL_469495|20 ···················· ···················· ···················· ···················· ···················· ····················
D 9599 hCoV-19/England/BRIS-12C595/2020|EPI_ISL_469497|20 ···················· ···················· ···················· ···················· ···················· ····················
D 9600 hCoV-19/England/BRIS-12C7BD/2020|EPI_ISL_469498|20 ···················· ···················· ···················· ···················· ···················· ····················
D 9601 hCoV-19/England/BRIS-12C744/2020|EPI_ISL_469499|20 ···················· ···················· ···················· ···················· ···················· ····················
D 9602 hCoV-19/England/CAMB-803F0/2020|EPI_ISL_470400|202 ···················· ···················· ···················· ···················· ···················· ····················
D 9603 hCoV-19/England/CAMB-7F7F8/2020|EPI_ISL_470401|202 ···················· ···················· ···················· ···················· ···················· ····················
D 9604 hCoV-19/England/CAMB-7F831/2020|EPI_ISL_470402|202 ···················· ···················· ···················· ···················· ···················· ····················
D 9605 hCoV-19/England/CAMB-80114/2020|EPI_ISL_470411|202 ···················· ···················· ···················· ···················· ···················· ····················
D 9606 hCoV-19/England/CAMB-7FA3B/2020|EPI_ISL_470413|202 ···················· ···················· ···················· ···················· ···················· ····················
D 9607 hCoV-19/England/CAMB-805AF/2020|EPI_ISL_470416|202 ···················· ···················· ···················· ···················· ···················· ····················
D 9608 hCoV-19/England/CAMB-80642/2020|EPI_ISL_470417|202 ···················· ···················· ···················· ···················· ···················· ····················
D 9609 hCoV-19/England/CAMB-8017E/2020|EPI_ISL_470418|202 ···················· ···················· ···················· ···················· ···················· ····················
D 9610 hCoV-19/England/CAMB-7DEB9/2020|EPI_ISL_470419|202 ···················· ···················· ···················· ···················· ···················· ····················
D 9611 hCoV-19/England/CAMB-800EA/2020|EPI_ISL_470424|202 ···················· ·A·················· ···················· ···················· ···················· ····················
D 9612 hCoV-19/England/CAMB-80581/2020|EPI_ISL_470425|202 ···················· ···················· ···················· ···················· ···················· ····················
D 9613 hCoV-19/England/CAMB-803E1/2020|EPI_ISL_470426|202 ···················· ···················· ···················· ···················· ···················· ····················
D 9614 hCoV-19/England/CAMB-80071/2020|EPI_ISL_470427|202 ···················· ···················· ···················· ···················· ···················· ····················
D 9615 hCoV-19/England/CAMB-805DC/2020|EPI_ISL_470430|202 ···················· ···················· ···················· ···················· ···················· ····················
D 9616 hCoV-19/England/CAMB-7FD41/2020|EPI_ISL_470431|202 ···················· ···················· ···················· ···················· ···················· ····················
D 9617 hCoV-19/England/CAMB-7FAB3/2020|EPI_ISL_470432|202 ···················· ···················· ···················· ···················· ···················· ····················
D 9618 hCoV-19/England/CAMB-7F664/2020|EPI_ISL_470433|202 ···················· ···················· ···················· ···················· ···················· ····················
D 9619 hCoV-19/England/CAMB-80466/2020|EPI_ISL_470434|202 ···················· ···················· ···················· ···················· ···················· ····················
D 9620 hCoV-19/England/CAMB-7F673/2020|EPI_ISL_470437|202 ···················· ···················· ···················· ···················· ···················· ····················
D 9621 hCoV-19/England/CAMB-805EB/2020|EPI_ISL_470438|202 ···················· ···················· ···················· ···················· ···················· ····················
D 9622 hCoV-19/England/CAMB-7F549/2020|EPI_ISL_470439|202 ···················· ···················· ···················· ···················· ···················· ····················
D 9623 hCoV-19/England/CAMB-80563/2020|EPI_ISL_470440|202 ···················· ···················· ···················· ···················· ···················· ····················
D 9624 hCoV-19/England/CAMB-7BFC7/2020|EPI_ISL_470441|202 ···················· ···················· ···················· ···················· ···················· ····················
D 9625 hCoV-19/England/CAMB-8067F/2020|EPI_ISL_470442|202 ···················· ···················· ···················· ···················· ···················· ····················
D 9626 hCoV-19/England/CAMB-8042A/2020|EPI_ISL_470443|202 ···················· ···················· ···················· ···················· ···················· ····················
D 9627 hCoV-19/England/CAMB-7F95C/2020|EPI_ISL_470445|202 ···················· ···················· ···················· ···················· ···················· ····················
D 9628 hCoV-19/England/CAMB-7DDBC/2020|EPI_ISL_470446|202 ···················· ···················· ···················· ···················· ···················· ····················
D 9629 hCoV-19/England/CAMB-80A73/2020|EPI_ISL_470447|202 ···················· ···················· ···················· ···················· ···················· ····················
D 9630 hCoV-19/England/CAMB-7FA95/2020|EPI_ISL_470449|202 ···················· ···················· ···················· ···················· ···················· ····················
D 9631 hCoV-19/England/CAMB-7F6FB/2020|EPI_ISL_470451|202 ···················· ···················· ···················· ···················· ···················· ····················
D 9632 hCoV-19/England/CAMB-7DF5C/2020|EPI_ISL_470452|202 ···················· ·A·················· ···················· ···················· ···················· ····················
D 9633 hCoV-19/England/CAMB-7F655/2020|EPI_ISL_470453|202 ···················· ···················· ···················· ···················· ···················· ····················

| | | | | | | | | | |
|---|---|---|---|---|---|---|---|---|---|
| D | 9634 | hCoV-19/England/CAMB-804FD/2020\|EPI_ISL_470454\|202 | ···················· | ···················· | ···················· | ···················· | ···················· | ···················· | |
| D | 9635 | hCoV-19/England/CAMB-8077C/2020\|EPI_ISL_470455\|202 | ···················· | ···················· | ···················· | ···················· | ···················· | ···················· | |
| D | 9636 | hCoV-19/England/CAMB-80545/2020\|EPI_ISL_470456\|202 | ···················· | ···················· | ···················· | ···················· | ···················· | ···················· | |
| D | 9637 | hCoV-19/England/CAMB-80536/2020\|EPI_ISL_470457\|202 | ···················· | ···················· | ···················· | ···················· | ···················· | ···················· | |
| D | 9638 | hCoV-19/England/CAMB-7DF7A/2020\|EPI_ISL_470459\|202 | ···················· | ···················· | ···················· | ···················· | ···················· | ···················· | |
| D | 9639 | hCoV-19/England/CAMB-7BF4F/2020\|EPI_ISL_470460\|202 | ···················· | ···················· | ···················· | ···················· | ···················· | ···················· | |
| D | 9640 | hCoV-19/England/CAMB-80457/2020\|EPI_ISL_470461\|202 | ···················· | ···················· | ···················· | ···················· | ···················· | ···················· | |
| D | 9641 | hCoV-19/England/CAMB-7F96B/2020\|EPI_ISL_470462\|202 | ···················· | ···················· | ···················· | ···················· | ···················· | ···················· | |
| D | 9642 | hCoV-19/England/CAMB-7F682/2020\|EPI_ISL_470464\|202 | ···················· | ···················· | ···················· | ···················· | ···················· | ···················· | |
| D | 9643 | hCoV-19/England/CAMB-7DE13/2020\|EPI_ISL_470465\|202 | ···················· | ···················· | ···················· | ···················· | ···················· | ···················· | |
| D | 9644 | hCoV-19/England/CAMB-801D8/2020\|EPI_ISL_470466\|202 | ···················· | ···················· | ···················· | ···················· | ···················· | ···················· | |
| D | 9645 | hCoV-19/England/CAMB-80518/2020\|EPI_ISL_470468\|202 | ···················· | ···················· | ···················· | ···················· | ···················· | ···················· | |
| D | 9646 | hCoV-19/England/CAMB-7BEAC/2020\|EPI_ISL_470471\|202 | ···················· | ···················· | ···················· | ···················· | ···················· | ···················· | |
| D | 9647 | hCoV-19/England/CAMB-7FAE0/2020\|EPI_ISL_470472\|202 | ···················· | ···················· | ···················· | ···················· | ···················· | ···················· | |
| D | 9648 | hCoV-19/England/CAMB-7F691/2020\|EPI_ISL_470473\|202 | ···················· | ···················· | ···················· | ···················· | ···················· | ···················· | |
| D | 9649 | hCoV-19/England/CAMB-80527/2020\|EPI_ISL_470474\|202 | ···················· | ···················· | ···················· | ···················· | ···················· | ···················· | |
| D | 9650 | hCoV-19/England/CAMB-8075E/2020\|EPI_ISL_470477\|202 | ···················· | ···················· | ···················· | ···················· | ···················· | ···················· | |
| D | 9651 | hCoV-19/England/CAMB-7F85F/2020\|EPI_ISL_470478\|202 | ···················· | ···················· | ···················· | ···················· | ···················· | ···················· | |
| D | 9652 | hCoV-19/England/CAMB-7F567/2020\|EPI_ISL_470479\|202 | ···················· | ···················· | ···················· | ···················· | ···················· | ···················· | |
| D | 9653 | hCoV-19/England/CAMB-7F5EF/2020\|EPI_ISL_470480\|202 | ···················· | ···················· | ···················· | ···················· | ···················· | ···················· | |
| D | 9654 | hCoV-19/England/CAMB-7DED7/2020\|EPI_ISL_470481\|202 | ···················· | ···················· | ···················· | ···················· | ···················· | ···················· | |
| D | 9655 | hCoV-19/England/CAMB-80387/2020\|EPI_ISL_470482\|202 | ···················· | ···················· | ···················· | ···················· | ···················· | ···················· | |
| D | 9656 | hCoV-19/England/CAMB-80141/2020\|EPI_ISL_470483\|202 | ···················· | ···················· | ···················· | ···················· | ···················· | ···················· | |
| D | 9657 | hCoV-19/England/CAMB-7DF98/2020\|EPI_ISL_470485\|202 | ···················· | ···················· | ···················· | ···················· | ···················· | ···················· | |
| D | 9658 | hCoV-19/England/CAMB-7FA59/2020\|EPI_ISL_470486\|202 | ···················· | ···················· | ···················· | ···················· | ···················· | ···················· | |
| D | 9659 | hCoV-19/England/CAMB-7F585/2020\|EPI_ISL_470487\|202 | ···················· | ···················· | ···················· | ···················· | ···················· | ···················· | |
| D | 9660 | hCoV-19/England/CAMB-807B8/2020\|EPI_ISL_470488\|202 | ···················· | ···················· | ···················· | ···················· | ···················· | ···················· | |
| D | 9661 | hCoV-19/England/CAMB-7F734/2020\|EPI_ISL_470490\|202 | ···················· | ···················· | ···················· | ···················· | ···················· | ···················· | |
| D | 9662 | hCoV-19/England/CAMB-7DD9E/2020\|EPI_ISL_470491\|202 | ···················· | ···················· | ···················· | ···················· | ···················· | ···················· | |
| D | 9663 | hCoV-19/England/CAMB-7FA0E/2020\|EPI_ISL_470495\|202 | ···················· | ···················· | ···················· | ···················· | ···················· | ···················· | |
| D | 9664 | hCoV-19/England/CAMB-7FF0F/2020\|EPI_ISL_470496\|202 | ···················· | ···················· | ···················· | ···················· | ···················· | ···················· | |
| D | 9665 | hCoV-19/England/CAMB-7BE60/2020\|EPI_ISL_470497\|202 | ···················· | ···················· | ···················· | ···················· | ···················· | ···················· | |
| D | 9666 | hCoV-19/England/CAMB-7F716/2020\|EPI_ISL_470498\|202 | ···················· | ···················· | ···················· | ···················· | ···················· | ···················· | |
| D | 9667 | hCoV-19/England/CAMB-805FA/2020\|EPI_ISL_470499\|202 | ···················· | ···················· | ···················· | ···················· | ···················· | ···················· | |
| D | 9668 | hCoV-19/USA/MI-MDHHS-SC20677/2020\|EPI_ISL_471701\|2 | ···················· | ···················· | ···················· | ···················· | ···················· | ···················· | |
| D | 9669 | hCoV-19/USA/MI-MDHHS-SC20678/2020\|EPI_ISL_471702\|2 | ···················· | ···················· | ···················· | ···················· | ···················· | ···················· | |
| D | 9670 | hCoV-19/USA/MI-MDHHS-SC20680/2020\|EPI_ISL_471704\|2 | ···················· | ···················· | ···················· | ···················· | ···················· | ···················· | |
| D | 9671 | hCoV-19/USA/MI-MDHHS-SC20683/2020\|EPI_ISL_471707\|2 | ···················· | ···················· | ···················· | ···················· | ···················· | ···················· | |
| D | 9673 | hCoV-19/USA/MI-MDHHS-SC20685/2020\|EPI_ISL_471709\|2 | ···················· | ···················· | ···················· | ···················· | ···················· | ···················· | |
| D | 9674 | hCoV-19/USA/MI-MDHHS-SC20688/2020\|EPI_ISL_471712\|2 | ···················· | ···················· | ···················· | ···················· | ···················· | ···················· | |
| D | 9675 | hCoV-19/USA/MI-MDHHS-SC20689/2020\|EPI_ISL_471713\|2 | ···················· | ···················· | ···················· | ···················· | ···················· | ···················· | |
| D | 9676 | hCoV-19/USA/MI-MDHHS-SC20690/2020\|EPI_ISL_471714\|2 | ···················· | ···················· | ···················· | ···················· | ···················· | ···················· | |
| D | 9677 | hCoV-19/USA/MI-MDHHS-SC20691/2020\|EPI_ISL_471715\|2 | ···················· | ···················· | ···················· | ···················· | ···················· | ···················· | |
| D | 9678 | hCoV-19/USA/MI-MDHHS-SC20692/2020\|EPI_ISL_471716\|2 | ···················· | ···················· | ···················· | ···················· | ···················· | ···················· | |
| D | 9679 | hCoV-19/USA/MI-MDHHS-SC20693/2020\|EPI_ISL_471717\|2 | ···················· | ···················· | ···················· | ···················· | ···················· | ···················· | |
| D | 9680 | hCoV-19/USA/MI-MDHHS-SC20696/2020\|EPI_ISL_471720\|2 | ···················· | ···················· | ···················· | ···················· | ···················· | ···················· | |
| D | 9681 | hCoV-19/USA/MI-MDHHS-SC20697/2020\|EPI_ISL_471721\|2 | ···················· | ···················· | ···················· | ···················· | ···················· | ···················· | |
| D | 9682 | hCoV-19/USA/MI-MDHHS-SC20698/2020\|EPI_ISL_471722\|2 | ···················· | ···················· | ···················· | ···················· | ···················· | ···················· | |
| D | 9683 | hCoV-19/USA/MI-MDHHS-SC20700/2020\|EPI_ISL_471724\|2 | ···················· | ···················· | ···················· | ···················· | ···················· | ···················· | |
| D | 9684 | hCoV-19/USA/MI-MDHHS-SC20701/2020\|EPI_ISL_471725\|2 | ···················· | ···················· | ···················· | ···················· | ···················· | ···················· | |
| D | 9685 | hCoV-19/USA/MI-MDHHS-SC20704/2020\|EPI_ISL_471728\|2 | ···················· | ···················· | ···················· | ···················· | ···················· | ···················· | |
| D | 9686 | hCoV-19/USA/MI-MDHHS-SC20705/2020\|EPI_ISL_471729\|2 | ···················· | ···················· | ···················· | ···················· | ···················· | ···················· | |
| D | 9687 | hCoV-19/USA/MI-MDHHS-SC20707/2020\|EPI_ISL_471731\|2 | ···················· | ···················· | ···················· | ···················· | ···················· | ···················· | |
| D | 9688 | hCoV-19/USA/MI-MDHHS-SC20708/2020\|EPI_ISL_471732\|2 | ···················· | ···················· | ···················· | ···················· | ···················· | ···················· | |
| D | 9689 | hCoV-19/USA/MI-MDHHS-SC20716/2020\|EPI_ISL_471740\|2 | ···················· | ···················· | ···················· | ···················· | ···················· | ···················· | |
| D | 9690 | hCoV-19/USA/MI-MDHHS-SC20718/2020\|EPI_ISL_471742\|2 | ···················· | ···················· | ···················· | ···················· | ···················· | ···················· | |
| D | 9691 | hCoV-19/USA/MI-MDHHS-SC20719/2020\|EPI_ISL_471743\|2 | ···················· | ···················· | ···················· | ···················· | ···················· | ···················· | |
| D | 9692 | hCoV-19/USA/MI-MDHHS-SC20721/2020\|EPI_ISL_471745\|2 | ···················· | ···················· | ···················· | ···················· | ···················· | ···················· | |
| D | 9693 | hCoV-19/USA/MI-MDHHS-SC20725/2020\|EPI_ISL_471749\|2 | ···················· | ···················· | ···················· | ···················· | ···················· | ···················· | |
| D | 9694 | hCoV-19/USA/MI-MDHHS-SC20726/2020\|EPI_ISL_471750\|2 | ···················· | ···················· | ···················· | ···················· | ···················· | ···················· | |
| D | 9695 | hCoV-19/USA/MI-MDHHS-SC20730/2020\|EPI_ISL_471754\|2 | ···················· | ···················· | ···················· | ···················· | ···················· | ···················· | |
| D | 9696 | hCoV-19/USA/MI-MDHHS-SC20746/2020\|EPI_ISL_471767\|2 | ···················· | ···················· | ···················· | ···················· | ···················· | ···················· | |
| D | 9697 | hCoV-19/USA/MI-MDHHS-SC20747/2020\|EPI_ISL_471768\|2 | ···················· | ···················· | ···················· | ···················· | ···················· | ···················· | |
| D | 9698 | hCoV-19/USA/MI-MDHHS-SC20748/2020\|EPI_ISL_471769\|2 | ···················· | ···················· | ···················· | ···················· | ···················· | ···················· | |
| D | 9699 | hCoV-19/USA/MI-MDHHS-SC20749/2020\|EPI_ISL_471770\|2 | ···················· | ···················R | ···················· | ···················· | ···················· | ···················· | 1 |
| D | 9700 | hCoV-19/USA/MI-MDHHS-SC20750/2020\|EPI_ISL_471771\|2 | ···················· | ···················· | ···················· | ···················· | ···················· | ···················· | |
| D | 9701 | hCoV-19/USA/MI-MDHHS-SC20751/2020\|EPI_ISL_471772\|2 | ···················· | ···················· | ···················· | ···················· | ···················· | ···················· | |
| D | 9702 | hCoV-19/USA/MI-MDHHS-SC20766/2020\|EPI_ISL_471787\|2 | ···················· | ···················· | ···················· | ···················· | ···················· | ···················· | |
| D | 9703 | hCoV-19/USA/MI-MDHHS-SC20767/2020\|EPI_ISL_471788\|2 | ···················· | ···················· | ···················· | ···················· | ···················· | ···················· | |
| D | 9704 | hCoV-19/USA/MI-MDHHS-SC20768/2020\|EPI_ISL_471789\|2 | ···················· | ···················· | ···················· | ···················· | ···················· | ···················· | |
| D | 9705 | hCoV-19/USA/MI-MDHHS-SC20769/2020\|EPI_ISL_471790\|2 | ···················· | ···A················ | ···················· | ···················· | ···················· | ···················· | |
| D | 9706 | hCoV-19/USA/MI-MDHHS-SC20770/2020\|EPI_ISL_471791\|2 | ···················· | ···················· | ···················· | ···················· | ···················· | ···················· | |
| D | 9707 | hCoV-19/USA/MI-MDHHS-SC20771/2020\|EPI_ISL_471792\|2 | ···················· | ···················· | ···················· | ···················· | ···················· | ···················· | |
| D | 9708 | hCoV-19/USA/MI-MDHHS-SC20772/2020\|EPI_ISL_471793\|2 | ···················· | ···················· | ···················· | ···················· | ···················· | ···················· | |
| D | 9709 | hCoV-19/USA/MI-MDHHS-SC20773/2020\|EPI_ISL_471794\|2 | ···················· | ···················· | ···················· | ···················· | ···················· | ···················· | |
| D | 9710 | hCoV-19/USA/MI-MDHHS-SC20774/2020\|EPI_ISL_471795\|2 | ···················· | ···················· | ···················· | ···················· | ···················· | ···················· | |
| D | 9711 | hCoV-19/USA/MI-MDHHS-SC20775/2020\|EPI_ISL_471796\|2 | ···················· | ···················· | ···················· | ···················· | ···················· | ···················· | |
| D | 9712 | hCoV-19/USA/MI-MDHHS-SC20776/2020\|EPI_ISL_471797\|2 | ···················· | ···················· | ···················· | ···················· | ···················· | ···················· | |

<table>
<tr><td>D</td><td>9713</td><td>hCoV-19/Scotland/CVR2319/2020|EPI_ISL_477901|2020-</td><td>····················</td><td>····················</td><td>····················</td><td>····················</td><td>····················</td><td>····················</td><td></td></tr>
<tr><td>D</td><td>9714</td><td>hCoV-19/Scotland/CVR2322/2020|EPI_ISL_477902|2020-</td><td>····················</td><td>···················A</td><td>····················</td><td>····················</td><td>····················</td><td>····················</td><td>1</td></tr>
<tr><td>D</td><td>9715</td><td>hCoV-19/Scotland/CVR2326/2020|EPI_ISL_477903|2020-</td><td>····················</td><td>···················A</td><td>····················</td><td>····················</td><td>····················</td><td>····················</td><td>1</td></tr>
<tr><td>D</td><td>9716</td><td>hCoV-19/Scotland/CVR2327/2020|EPI_ISL_477904|2020-</td><td>····················</td><td>···················A</td><td>····················</td><td>····················</td><td>····················</td><td>····················</td><td>1</td></tr>
<tr><td>D</td><td>9717</td><td>hCoV-19/Scotland/CVR2330/2020|EPI_ISL_477905|2020-</td><td>····················</td><td>···················A</td><td>····················</td><td>····················</td><td>····················</td><td>····················</td><td>1</td></tr>
<tr><td>D</td><td>9718</td><td>hCoV-19/Scotland/CVR2331/2020|EPI_ISL_477906|2020-</td><td>····················</td><td>····················</td><td>····················</td><td>····················</td><td>····················</td><td>····················</td><td></td></tr>
<tr><td>D</td><td>9719</td><td>hCoV-19/Scotland/CVR2337/2020|EPI_ISL_477907|2020-</td><td>····················</td><td>····················</td><td>····················</td><td>····················</td><td>····················</td><td>····················</td><td></td></tr>
<tr><td>D</td><td>9720</td><td>hCoV-19/Scotland/CVR2339/2020|EPI_ISL_477909|2020-</td><td>····················</td><td>····················</td><td>····················</td><td>····················</td><td>····················</td><td>····················</td><td></td></tr>
<tr><td>D</td><td>9721</td><td>hCoV-19/Scotland/CVR2342/2020|EPI_ISL_477910|2020-</td><td>····················</td><td>····················</td><td>····················</td><td>····················</td><td>····················</td><td>····················</td><td></td></tr>
<tr><td>D</td><td>9722</td><td>hCoV-19/Scotland/CVR2348/2020|EPI_ISL_477911|2020-</td><td>····················</td><td>····················</td><td>····················</td><td>····················</td><td>····················</td><td>····················</td><td></td></tr>
<tr><td>D</td><td>9723</td><td>hCoV-19/Scotland/CVR2353/2020|EPI_ISL_477912|2020-</td><td>····················</td><td>····················</td><td>····················</td><td>····················</td><td>····················</td><td>····················</td><td></td></tr>
<tr><td>D</td><td>9724</td><td>hCoV-19/Scotland/CVR2355/2020|EPI_ISL_477914|2020-</td><td>····················</td><td>···················A</td><td>····················</td><td>····················</td><td>····················</td><td>····················</td><td>1</td></tr>
<tr><td>D</td><td>9725</td><td>hCoV-19/Scotland/CVR2356/2020|EPI_ISL_477915|2020-</td><td>····················</td><td>····················</td><td>····················</td><td>····················</td><td>····················</td><td>····················</td><td></td></tr>
<tr><td>D</td><td>9726</td><td>hCoV-19/Scotland/CVR2358/2020|EPI_ISL_477916|2020-</td><td>····················</td><td>··········T········A</td><td>····················</td><td>····················</td><td>····················</td><td>····················</td><td>1</td></tr>
<tr><td>D</td><td>9727</td><td>hCoV-19/Scotland/CVR2363/2020|EPI_ISL_477918|2020-</td><td>····················</td><td>····················</td><td>····················</td><td>····················</td><td>····················</td><td>····················</td><td></td></tr>
<tr><td>D</td><td>9728</td><td>hCoV-19/Scotland/CVR2370/2020|EPI_ISL_477919|2020-</td><td>····················</td><td>····················</td><td>····················</td><td>····················</td><td>····················</td><td>····················</td><td></td></tr>
<tr><td>D</td><td>9729</td><td>hCoV-19/USA/OR-OHSU-0126/2020|EPI_ISL_468105|2020-</td><td>····················</td><td>····················</td><td>····················</td><td>····················</td><td>····················</td><td>····················</td><td></td></tr>
<tr><td>D</td><td>9730</td><td>hCoV-19/England/BRIS-12BDBC/2020|EPI_ISL_469427|20</td><td>····················</td><td>····················</td><td>····················</td><td>····················</td><td>····················</td><td>····················</td><td></td></tr>
<tr><td>D</td><td>9731</td><td>hCoV-19/England/BRIS-12BF6B/2020|EPI_ISL_469428|20</td><td>····················</td><td>····················</td><td>····················</td><td>····················</td><td>····················</td><td>····················</td><td></td></tr>
<tr><td>D</td><td>9732</td><td>hCoV-19/England/BRIS-12C3AA/2020|EPI_ISL_469434|20</td><td>····················</td><td>····················</td><td>····················</td><td>····················</td><td>····················</td><td>····················</td><td></td></tr>
<tr><td>D</td><td>9733</td><td>hCoV-19/England/BRIS-12B912/2020|EPI_ISL_469435|20</td><td>····················</td><td>····················</td><td>····················</td><td>····················</td><td>····················</td><td>····················</td><td></td></tr>
<tr><td>D</td><td>9734</td><td>hCoV-19/England/CAMB-7BE7F/2020|EPI_ISL_470403|202</td><td>····················</td><td>····················</td><td>····················</td><td>····················</td><td>····················</td><td>····················</td><td></td></tr>
<tr><td>D</td><td>9735</td><td>hCoV-19/England/CAMB-8041B/2020|EPI_ISL_470404|202</td><td>····················</td><td>····················</td><td>····················</td><td>····················</td><td>····················</td><td>····················</td><td></td></tr>
<tr><td>D</td><td>9736</td><td>hCoV-19/England/CAMB-7F78F/2020|EPI_ISL_470405|202</td><td>····················</td><td>····················</td><td>····················</td><td>····················</td><td>····················</td><td>····················</td><td></td></tr>
<tr><td>D</td><td>9737</td><td>hCoV-19/England/CAMB-7F9C5/2020|EPI_ISL_470406|202</td><td>····················</td><td>····················</td><td>····················</td><td>····················</td><td>····················</td><td>····················</td><td></td></tr>
<tr><td>D</td><td>9738</td><td>hCoV-19/England/CAMB-800BD/2020|EPI_ISL_470407|202</td><td>····················</td><td>····················</td><td>····················</td><td>····················</td><td>····················</td><td>····················</td><td></td></tr>
<tr><td>D</td><td>9739</td><td>hCoV-19/England/CAMB-7F7DA/2020|EPI_ISL_470408|202</td><td>····················</td><td>····················</td><td>····················</td><td>····················</td><td>····················</td><td>····················</td><td></td></tr>
<tr><td>D</td><td>9740</td><td>hCoV-19/England/CAMB-80150/2020|EPI_ISL_470409|202</td><td>····················</td><td>····················</td><td>····················</td><td>····················</td><td>····················</td><td>····················</td><td></td></tr>
<tr><td>D</td><td>9741</td><td>hCoV-19/USA/MI-MDHHS-SC20715/2020|EPI_ISL_471739|2</td><td>····················</td><td>····················</td><td>····················</td><td>····················</td><td>····················</td><td>····················</td><td></td></tr>
</table>

Group 2 - Jun 2020- Sep 2020

| A | B | C | D | E | F | G | H | I | J | K | L |
|---|---|---|---|---|---|---|---|---|---|---|---|
| Database | Database ID | GISAID ID | GCTGCAATCGTGCTACAACT | TGAACTGTTGCGACTACGTG | TTCGGAAGAGACAGGTACGTT | CACACAATCGAAGCGCAGTA | GCTGGTGCTGCAGCTTATTA | AGGGTCAAGTGCACAGTCTA | mismatches? | mismatches? | primer set? |
| E | 1 | hCoV-19/USA/WI-UW-418/2020\|EPI_ISL_480400\|2020-04- | ···················· | ···················· | ···················· | ···················· | ··············N····· | ·······N·····N······ | | | |
| E | 2 | hCoV-19/USA/WI-UW-419/2020\|EPI_ISL_480401\|2020-04- | ···················· | ···················· | ···················· | ···················· | ···················· | ···················· | | | |
| E | 3 | hCoV-19/USA/WI-UW-420/2020\|EPI_ISL_480402\|2020-04- | ···················· | ···················· | ···················· | ···················· | ···················· | ···················· | | | |
| E | 4 | hCoV-19/USA/WI-UW-421/2020\|EPI_ISL_480403\|2020-04- | ···················· | ···················· | ···················· | ···················· | ···················· | ···················· | | | |
| E | 5 | hCoV-19/USA/WI-UW-422/2020\|EPI_ISL_480404\|2020-04- | ···················· | ···················· | ···················· | ···················· | ···················· | ···················· | | | |
| E | 6 | hCoV-19/USA/WI-UW-424/2020\|EPI_ISL_480406\|2020-04- | ···················· | ···················· | ···················· | ···················· | ···················· | ···················· | | | |
| E | 7 | hCoV-19/USA/WI-UW-425/2020\|EPI_ISL_480407\|2020-04- | ···················· | ···················· | ···················· | ···················· | ···················· | ···················· | | | |
| E | 8 | hCoV-19/USA/WI-UW-426/2020\|EPI_ISL_480408\|2020-04- | ···················· | ···················· | ···················· | ···················· | ···················· | ···················· | | | |
| E | 9 | hCoV-19/USA/WI-UW-427/2020\|EPI_ISL_480409\|2020-04- | ···················· | ···················· | ···················· | ···················· | ···················· | ···················· | | | |
| E | 10 | hCoV-19/Finland/5May46S12/2020\|EPI_ISL_481700\|2020 | ···················· | ···················· | ···················· | ···················· | ···················· | ···················· | | | |
| E | 11 | hCoV-19/Finland/5May47S12/2020\|EPI_ISL_481701\|2020 | ···················· | ···················· | ···················· | ···················· | ···················· | ···················· | | | |
| E | 12 | hCoV-19/Finland/5May49S12/2020\|EPI_ISL_481702\|2020 | ···················· | ···················· | ···················· | ···················· | ···················· | ···················· | | | |
| E | 13 | hCoV-19/Finland/5May49S6/2020\|EPI_ISL_481703\|2020- | ···················· | ···················· | ···················· | ···················· | ···················· | ···················· | | | |
| E | 14 | hCoV-19/Finland/5May56S6/2020\|EPI_ISL_481704\|2020- | ···················· | ···················· | ···················· | ···················· | ···················· | ···················· | | | |
| E | 15 | hCoV-19/Finland/5May74S2/2020\|EPI_ISL_481705\|2020- | ···················· | ···················· | ···················· | ···················· | ···················· | ···················· | | | |
| E | 16 | hCoV-19/Finland/5May84S2/2020\|EPI_ISL_481706\|2020- | ···················· | ···················· | ···················· | ···················· | ···················· | ···················· | | | |
| E | 17 | hCoV-19/Finland/5S54S2/2020\|EPI_ISL_481707\|2020-04 | ···················· | ···················· | ···················· | ···················· | ···················· | ···················· | | | |
| E | 18 | hCoV-19/Finland/6A928SIIB/2020\|EPI_ISL_481708\|2020 | ···················· | ···················· | ···················· | ···················· | ···················· | ···················· | | | |
| E | 19 | hCoV-19/Finland/6A937SIIB/2020\|EPI_ISL_481709\|2020 | ···················· | ···················· | ···················· | ···················· | ···················· | ···················· | | | |
| E | 20 | hCoV-19/Finland/6A953SIIB/2020\|EPI_ISL_481710\|2020 | ···················· | ···················· | ···················· | ···················· | ···················· | ···················· | | | |
| E | 21 | hCoV-19/Finland/6A959SIIC/2020\|EPI_ISL_481711\|2020 | ···················· | ···················· | ···················· | ···················· | ···················· | ···················· | | | |
| E | 22 | hCoV-19/Finland/6A962SIIC/2020\|EPI_ISL_481712\|2020 | ···················· | ···················· | ···················· | ···················· | ···················· | ···················· | | | |
| E | 23 | hCoV-19/Finland/6A973SIIC/2020\|EPI_ISL_481713\|2020 | ···················· | ···················· | ···················· | ···················· | ···················· | ···················· | | | |
| E | 24 | hCoV-19/Finland/6May47S11/2020\|EPI_ISL_481714\|2020 | ···················· | ···················· | ···················· | ···················· | ···················· | ···················· | | | |
| E | 25 | hCoV-19/Finland/7May68S22/2020\|EPI_ISL_481715\|2020 | ···················· | ···················· | ···················· | ···················· | ···················· | ···················· | | | |
| E | 26 | hCoV-19/Finland/7May75S23/2020\|EPI_ISL_481717\|2020 | ···················· | ···················· | ···················· | ···················· | ···················· | ···················· | | | |
| E | 27 | hCoV-19/Finland/7May92S23/2020\|EPI_ISL_481718\|2020 | ···················· | ···················· | ···················· | ···················· | ···················· | ···················· | | | |
| E | 28 | hCoV-19/Finland/8May11S7/2020\|EPI_ISL_481719\|2020- | ···················· | ···················· | ···················· | ···················· | ···················· | ···················· | | | |
| E | 29 | hCoV-19/Finland/8May18S18/2020\|EPI_ISL_481720\|2020 | ···················· | ···················· | ···················· | ···················· | ···················· | ···················· | | | |
| E | 30 | hCoV-19/Finland/8May21S18/2020\|EPI_ISL_481721\|2020 | ···················· | ···················· | ···················· | ···················· | ···················· | ···················· | | | |
| E | 31 | hCoV-19/Finland/8May24S14/2020\|EPI_ISL_481722\|2020 | ···················· | ···················· | ···················· | ···················· | ···················· | ···················· | | | |
| E | 32 | hCoV-19/Finland/8May30S7/2020\|EPI_ISL_481723\|2020- | ···················· | ···················· | ···················· | ···················· | ···················· | ···················· | | | |
| E | 33 | hCoV-19/Finland/8May35S7/2020\|EPI_ISL_481724\|2020- | ···················· | ···················· | ···················· | ···················· | ···················· | ···················· | | | |
| E | 34 | hCoV-19/Finland/8May38S18/2020\|EPI_ISL_481725\|2020 | ···················· | ···················· | ···················· | ···················· | ···················· | ···················· | | | |
| E | 35 | hCoV-19/Finland/8May43S7/2020\|EPI_ISL_481726\|2020- | ···················· | ···················· | ···················· | ···················· | ···················· | ···················· | | | |
| E | 36 | hCoV-19/Finland/8May65S11/2020\|EPI_ISL_481727\|2020 | ···················· | ···················· | ···················· | ···················· | ···················· | ···················· | | | |
| E | 37 | hCoV-19/Finland/8May89S14/2020\|EPI_ISL_481729\|2020 | ···················· | ···················· | ···················· | ···················· | ···················· | ···················· | | | |
| E | 38 | hCoV-19/Finland/9A18S12/2020\|EPI_ISL_481730\|2020-0 | ···················· | ···················· | ···················· | ···················· | ···················· | ···················· | | | |
| E | 39 | hCoV-19/Finland/9A29S10/2020\|EPI_ISL_481731\|2020-0 | ···················· | ···················· | ···················· | ···················· | ···················· | ···················· | | | |
| E | 40 | hCoV-19/Finland/9A30S12/2020\|EPI_ISL_481732\|2020-0 | ···················· | ···················· | ···················· | ···················· | ···················· | ···················· | | | |
| E | 41 | hCoV-19/Finland/9A40S12/2020\|EPI_ISL_481733\|2020-0 | ···················· | ···················· | ···················· | ···················· | ···················· | ···················· | | | |
| E | 42 | hCoV-19/Finland/9A784S11A/2020\|EPI_ISL_481734\|2020 | ···················· | ···················· | ···················· | ···················· | ···················· | ···················· | | | |
| E | 43 | hCoV-19/Finland/9A785S11A/2020\|EPI_ISL_481735\|2020 | ···················· | ···················· | ···················· | ···················· | ···················· | ···················· | | | |
| E | 44 | hCoV-19/Finland/9A797S11A/2020\|EPI_ISL_481736\|2020 | ···················· | ···················· | ···················· | ···················· | ···················· | ···················· | | | |
| E | 45 | hCoV-19/Finland/9A798S11A/2020\|EPI_ISL_481737\|2020 | ···················· | ···················· | ···················· | ···················· | ···················· | ···················· | | | |
| E | 46 | hCoV-19/Finland/9A800S11A/2020\|EPI_ISL_481738\|2020 | ···················· | ···················· | ···················· | ···················· | ···················· | ···················· | | | |
| E | 47 | hCoV-19/Finland/9A81S12/2020\|EPI_ISL_481739\|2020-0 | ···················· | ···················· | ···················· | ···················· | ···················· | ···················· | | | |
| E | 48 | hCoV-19/USA/LA-GBCL-26556572/2020\|EPI_ISL_485200\|2 | ···················· | ···················· | ···················· | ···················· | ···················· | ···················· | | | |
| E | 49 | hCoV-19/USA/LA-GBCL-26556574/2020\|EPI_ISL_485201\|2 | ···················· | ···················· | ···················· | ···················· | ···················· | ···················· | | | |
| E | 50 | hCoV-19/USA/LA-GBCL-26556575/2020\|EPI_ISL_485202\|2 | ···················· | ···················· | ···················· | ···················· | ···················· | ···················· | | | |
| E | 51 | hCoV-19/USA/LA-GBCL-26556576/2020\|EPI_ISL_485203\|2 | ···················· | ···················· | ···················· | ···················· | ···················· | ···················· | | | |
| E | 52 | hCoV-19/USA/LA-GBCL-26556578/2020\|EPI_ISL_485204\|2 | ···················· | ···················· | ···················· | ···················· | ···················· | ···················· | | | |
| E | 53 | hCoV-19/USA/LA-GBCL-26556579/2020\|EPI_ISL_485205\|2 | ···················· | ···················· | ···················· | ···················· | ···················· | ···················· | | | |
| E | 54 | hCoV-19/USA/LA-GBCL-26556581/2020\|EPI_ISL_485206\|2 | ···················· | ···················· | ···················· | ···················· | ···················· | ···················· | | | |
| E | 55 | hCoV-19/USA/LA-GBCL-26556585/2020\|EPI_ISL_485207\|2 | ···················· | ···················· | ···················· | ···················· | ···················· | ···················· | | | |
| E | 56 | hCoV-19/USA/LA-GBCL-26556586/2020\|EPI_ISL_485208\|2 | ···················· | ···················· | ···················· | ···················· | ···················· | ···················· | | | |
| E | 57 | hCoV-19/USA/LA-GBCL-26556587/2020\|EPI_ISL_485209\|2 | ···················· | ···················· | ···················· | ···················· | ···················· | ···················· | | | |
| E | 58 | hCoV-19/USA/LA-GBCL-26556588/2020\|EPI_ISL_485210\|2 | ···················· | ···················· | ···················· | ···················· | ···················· | ···················· | | | |
| E | 59 | hCoV-19/USA/LA-GBCL-26629192/2020\|EPI_ISL_485214\|2 | ···················· | ···················· | ···················· | ···················· | ···················· | ···················· | | | |
| E | 60 | hCoV-19/USA/LA-GBCL-26629202/2020\|EPI_ISL_485217\|2 | ···················· | ···················· | ···················· | ···················· | ···················· | ···················· | | | |
| E | 61 | hCoV-19/USA/LA-GBCL-26629204/2020\|EPI_ISL_485218\|2 | ···················· | ···················· | ···················· | ···················· | ···················· | ···················· | | | |
| E | 62 | hCoV-19/USA/LA-GBCL-26629209/2020\|EPI_ISL_485219\|2 | ···················· | ···················· | ···················· | ···················· | ···················· | ···················· | | | |
| E | 63 | hCoV-19/USA/LA-GBCL-26629211/2020\|EPI_ISL_485220\|2 | ···················· | ···················· | ···················· | ···················· | ···················· | ···················· | | | |
| E | 64 | hCoV-19/USA/LA-GBCL-26629213/2020\|EPI_ISL_485221\|2 | ···················· | ···················· | ···················· | ···················· | ···················· | ···················· | | | |
| E | 65 | hCoV-19/USA/LA-GBCL-26629217/2020\|EPI_ISL_485222\|2 | ···················· | ···················· | ···················· | ···················· | ···················· | ···················· | | | |
| E | 66 | hCoV-19/USA/LA-GBCL-26629218/2020\|EPI_ISL_485223\|2 | ···················· | ···················· | ···················· | ···················· | ···················· | ···················· | | | |
| E | 67 | hCoV-19/USA/LA-GBCL-26629227/2020\|EPI_ISL_485226\|2 | ···················· | ···················· | ···················· | ···················· | ···················· | ···················· | | | |
| E | 68 | hCoV-19/USA/LA-GBCL-26629228/2020\|EPI_ISL_485227\|2 | ···················· | ···················· | ···················· | ···················· | ···················· | ···················· | | | |
| E | 69 | hCoV-19/USA/LA-GBCL-26629230/2020\|EPI_ISL_485228\|2 | ···················· | ···················· | ···················· | ···················· | ···················· | ···················· | | | |

| | | | | | | | | |
|---|---|---|---|---|---|---|---|---|
| E | 70 hCoV-19/USA/LA-GBCL-26629234/2020\|EPI_ISL_485230\|2 | ···················· | ···················· | ····················· | ···················· | ···················· | ···················· | |
| E | 71 hCoV-19/USA/LA-GBCL-26629237/2020\|EPI_ISL_485231\|2 | ···················· | ···················· | ····················· | ···················· | ···················· | ···················· | |
| E | 72 hCoV-19/USA/LA-GBCL-26629239/2020\|EPI_ISL_485232\|2 | ···················· | ···················· | ····················· | ···················· | ···················· | ···················· | |
| E | 73 hCoV-19/USA/LA-GBCL-26629240/2020\|EPI_ISL_485233\|2 | ···················· | ···················· | ····················· | ···················· | ···················· | ···················· | |
| E | 74 hCoV-19/USA/LA-GBCL-26629241/2020\|EPI_ISL_485234\|2 | ···················· | ···················· | ····················· | ···················· | ···················· | ···················· | |
| E | 75 hCoV-19/USA/LA-GBCL-26629247/2020\|EPI_ISL_485235\|2 | ···················· | ···················· | ····················· | ···················· | ···················· | ···················· | |
| E | 76 hCoV-19/USA/LA-GBCL-26629254/2020\|EPI_ISL_485237\|2 | ···················· | ···················· | ····················· | ···················· | ···················· | ···················· | |
| E | 77 hCoV-19/USA/LA-GBCL-26629255/2020\|EPI_ISL_485238\|2 | ···················· | ···················· | ····················· | ···················· | ···················· | ···················· | |
| E | 78 hCoV-19/USA/LA-GBCL-26629260/2020\|EPI_ISL_485240\|2 | ···················· | ···················· | ····················· | ···················· | ···················· | ···················· | |
| E | 79 hCoV-19/USA/LA-GBCL-26629267/2020\|EPI_ISL_485243\|2 | ···················· | ···················· | ····················· | ···················· | ···················· | ···················· | |
| E | 80 hCoV-19/USA/LA-GBCL-26629268/2020\|EPI_ISL_485244\|2 | ···················· | ···················· | ····················· | ···················· | ···················· | ···················· | |
| E | 81 hCoV-19/USA/LA-GBCL-26674720/2020\|EPI_ISL_485248\|2 | ···················· | ···················· | ····················· | ···················· | ···················· | ···················· | |
| E | 82 hCoV-19/USA/LA-GBCL-26674721/2020\|EPI_ISL_485249\|2 | ···················· | ···················· | ····················· | ···················· | ···················· | ···················· | |
| E | 83 hCoV-19/USA/LA-GBCL-26674722/2020\|EPI_ISL_485250\|2 | ···················· | ···················· | ····················· | ···················· | ···················· | ···················· | |
| E | 84 hCoV-19/USA/LA-GBCL-26674728/2020\|EPI_ISL_485254\|2 | ···················· | ···················· | ····················· | ···················· | ···················· | ···················· | |
| E | 85 hCoV-19/USA/LA-GBCL-26674732/2020\|EPI_ISL_485257\|2 | ···················· | ···················· | ····················· | ···················· | ···················· | ···················· | |
| E | 86 hCoV-19/USA/LA-GBCL-26674733/2020\|EPI_ISL_485258\|2 | ···················· | ···················· | ····················· | ···················· | ···················· | ···················· | |
| E | 87 hCoV-19/USA/LA-GBCL-26674735/2020\|EPI_ISL_485259\|2 | ···················· | ···················· | ····················· | ···················· | ···················· | ···················· | |
| E | 88 hCoV-19/USA/LA-GBCL-26684430/2020\|EPI_ISL_485260\|2 | ···················· | ···················· | ····················· | ···················· | ···················· | ···················· | |
| E | 89 hCoV-19/USA/LA-GBCL-26684437/2020\|EPI_ISL_485263\|2 | ···················· | ···················· | ····················· | ···················· | ···················· | ···················· | |
| E | 90 hCoV-19/USA/LA-GBCL-26684438/2020\|EPI_ISL_485264\|2 | ···················· | ···················· | ····················· | ···················· | ···················· | ···················· | |
| E | 91 hCoV-19/USA/LA-GBCL-26684440/2020\|EPI_ISL_485265\|2 | ···················· | ···················· | ····················· | ···················· | ···················· | ···················· | |
| E | 92 hCoV-19/USA/LA-GBCL-26684444/2020\|EPI_ISL_485269\|2 | ···················· | ···················· | ····················· | ···················· | ···················· | ···················· | |
| E | 93 hCoV-19/USA/LA-GBCL-26684446/2020\|EPI_ISL_485270\|2 | ···················· | ···················· | ····················· | ···················· | ···················· | ···················· | |
| E | 94 hCoV-19/USA/LA-GBCL-26684447/2020\|EPI_ISL_485271\|2 | ···················· | ···················· | ····················· | ···················· | ···················· | ···················· | |
| E | 95 hCoV-19/USA/LA-GBCL-26684453/2020\|EPI_ISL_485275\|2 | ···················· | ···················· | ····················· | ···················· | ···················· | ···················· | |
| E | 96 hCoV-19/USA/LA-GBCL-26684457/2020\|EPI_ISL_485278\|2 | ···················· | ···················· | ····················· | ···················· | ···················· | ···················· | |
| E | 97 hCoV-19/USA/LA-GBCL-26684458/2020\|EPI_ISL_485279\|2 | ···················· | ···················· | ····················· | ···················· | ···················· | ···················· | |
| E | 98 hCoV-19/USA/LA-GBCL-26684459/2020\|EPI_ISL_485280\|2 | ···················· | ···················· | ····················· | ···················· | ···················· | ···················· | |
| E | 99 hCoV-19/USA/LA-GBCL-26684460/2020\|EPI_ISL_485281\|2 | ···················· | ···················· | ····················· | ···················· | ···················· | ···················· | |
| E | 100 hCoV-19/USA/LA-GBCL-26684461/2020\|EPI_ISL_485282\|2 | ···················· | ···················· | ····················· | ···················· | ···················· | ···················· | |
| E | 101 hCoV-19/USA/LA-GBCL-26684462/2020\|EPI_ISL_485283\|2 | ···················· | ···················· | ····················· | ···················· | ···················· | ···················· | |
| E | 102 hCoV-19/USA/LA-GBCL-26684463/2020\|EPI_ISL_485284\|2 | ···················· | ···················· | ····················· | ···················· | ···················· | ···················· | |
| E | 103 hCoV-19/USA/LA-GBCL-26684466/2020\|EPI_ISL_485286\|2 | ···················· | ···················· | ····················· | ···················· | ···················· | ···················· | |
| E | 104 hCoV-19/USA/LA-GBCL-26684467/2020\|EPI_ISL_485287\|2 | ···················· | ···················· | ····················· | ···················· | ···················· | ···················· | |
| E | 105 hCoV-19/USA/LA-GBCL-26684470/2020\|EPI_ISL_485288\|2 | ···················· | ···················· | ····················· | ···················· | ···················· | ···················· | |
| E | 106 hCoV-19/USA/LA-GBCL-26684471/2020\|EPI_ISL_485289\|2 | ···················· | ···················· | ····················· | ···················· | ···················· | ···················· | |
| E | 107 hCoV-19/USA/LA-GBCL-26684472/2020\|EPI_ISL_485290\|2 | ···················· | ···················· | ····················· | ···················· | ···················· | ···················· | |
| E | 108 hCoV-19/USA/LA-GBCL-26684473/2020\|EPI_ISL_485291\|2 | ···················· | ···················· | ····················· | ···················· | ···················· | ···················· | |
| E | 109 hCoV-19/USA/LA-GBCL-26684478/2020\|EPI_ISL_485294\|2 | ···················· | ···················· | ····················· | ···················· | ···················· | ···················· | |
| E | 110 hCoV-19/USA/LA-GBCL-26684480/2020\|EPI_ISL_485295\|2 | ···················· | ···················· | ····················· | ···················· | ···················· | ···················· | |
| E | 111 hCoV-19/USA/LA-GBCL-26684481/2020\|EPI_ISL_485296\|2 | ···················· | ···················· | ····················· | ···················· | ···················· | ···················· | |
| E | 112 hCoV-19/USA/LA-GBCL-26684484/2020\|EPI_ISL_485297\|2 | ···················· | ···················· | ····················· | ···················· | ···················· | ···················· | |
| E | 113 hCoV-19/USA/LA-GBCL-26684485/2020\|EPI_ISL_485298\|2 | ···················· | ···················· | ····················· | ···················· | ···················· | ···················· | |
| E | 114 hCoV-19/Switzerland/BE-ETHZ-160017/2020\|EPI_ISL_48 | ···················· | ···················· | ····················· | ···················· | ···················· | ···················· | |
| E | 115 hCoV-19/Switzerland/ZH-ETHZ-160018/2020\|EPI_ISL_48 | ···················· | ···················· | ····················· | ···················· | ···················· | ···················· | |
| E | 116 hCoV-19/Switzerland/ZH-ETHZ-160019/2020\|EPI_ISL_48 | ···················· | ···················· | ····················· | ···················· | ·······S············ | ···················· | |
| E | 117 hCoV-19/Switzerland/ZH-ETHZ-160020/2020\|EPI_ISL_48 | ···················· | ···················· | ····················· | ···················· | ···················· | ···················· | |
| E | 118 hCoV-19/Switzerland/ZH-ETHZ-160021/2020\|EPI_ISL_48 | ···················· | ···················· | ····················· | ···················· | ···················· | ···················· | |
| E | 119 hCoV-19/Switzerland/GR-ETHZ-170000/2020\|EPI_ISL_48 | ···················· | ···················· | ····················· | ···················· | ···················· | ···················· | |
| E | 120 hCoV-19/Switzerland/SZ-ETHZ-170001/2020\|EPI_ISL_48 | ···················· | ···················· | ····················· | ···················· | ···················· | ···················· | |
| E | 121 hCoV-19/Switzerland/AG-ETHZ-170002/2020\|EPI_ISL_48 | ···················· | ···················· | ··················R·· | ···················· | ···················· | ···················· | 1 |
| E | 122 hCoV-19/Switzerland/BE-ETHZ-170003/2020\|EPI_ISL_48 | ···········T········ | ···················· | ····················· | ···················· | ···················· | ···················· | |
| E | 123 hCoV-19/Switzerland/UR-ETHZ-170004/2020\|EPI_ISL_48 | ···········T········ | ···················· | ····················· | ···················· | ···················· | ···················· | |
| E | 124 hCoV-19/Switzerland/BL-ETHZ-170007/2020\|EPI_ISL_48 | ···················· | ···················· | ····················· | ···················· | ···················· | ···················· | |
| E | 125 hCoV-19/Switzerland/ZH-ETHZ-170008/2020\|EPI_ISL_48 | ···················· | ···················· | ····················· | ···················· | ···················· | ···················· | |
| E | 126 hCoV-19/Switzerland/UR-ETHZ-170010/2020\|EPI_ISL_48 | ···········T········ | ···················· | ····················· | ···················· | ···················· | ···················· | |
| E | 127 hCoV-19/Switzerland/ZH-ETHZ-170011/2020\|EPI_ISL_48 | ···················· | ···················· | ····················· | ···················· | ···················· | ···················· | |
| E | 128 hCoV-19/Switzerland/BL-ETHZ-170013/2020\|EPI_ISL_48 | ···················· | ···················· | ····················· | ···················· | ···················· | ···················· | |
| E | 129 hCoV-19/Switzerland/AG-ETHZ-170014/2020\|EPI_ISL_48 | ···················· | ···················· | ····················· | ···················· | ···················· | ···················· | |
| E | 130 hCoV-19/Switzerland/BE-ETHZ-170016/2020\|EPI_ISL_48 | ···················· | ···················· | ····················· | ···················· | ·······S············ | ···················· | |
| E | 131 hCoV-19/Switzerland/ZH-ETHZ-170018/2020\|EPI_ISL_48 | ···················· | ···················· | ····················· | ···················· | ···················· | ···················· | |
| E | 132 hCoV-19/Switzerland/GR-ETHZ-170019/2020\|EPI_ISL_48 | ···················· | ···················· | ····················· | ···················· | ···················· | ···················· | |
| E | 133 hCoV-19/Switzerland/ZH-ETHZ-170020/2020\|EPI_ISL_48 | ···················· | ···················· | ····················· | ···················· | ···················· | ···················· | |
| E | 134 hCoV-19/Switzerland/VD-ETHZ-170021/2020\|EPI_ISL_48 | ···················· | ···················· | ····················· | ···················· | ···················· | ···················· | |
| E | 135 hCoV-19/Switzerland/SG-ETHZ-170022/2020\|EPI_ISL_48 | ···················· | ···················· | ····················· | ···················· | ···················· | ···················· | |
| E | 136 hCoV-19/Switzerland/BE-ETHZ-170023/2020\|EPI_ISL_48 | ···········T········ | ···················· | ····················· | ···················· | ···················· | ···················· | |
| E | 137 hCoV-19/Switzerland/BL-ETHZ-170025/2020\|EPI_ISL_48 | ···················· | ···················· | ····················· | ···················· | ···················· | ···················· | |
| E | 138 hCoV-19/Switzerland/BS-ETHZ-170026/2020\|EPI_ISL_48 | ···················· | ···················· | ····················· | ···················· | ···················· | ···················· | |
| E | 139 hCoV-19/Scotland/EDB5951/2020\|EPI_ISL_487800\|2020- | ···················· | ···················A | ····················· | ···················· | ···················· | ···················· | 1 |
| E | 140 hCoV-19/Scotland/EDB4252/2020\|EPI_ISL_487801\|2020- | ···················· | ···················A | ····················· | ···················· | ···················· | ···················· | 1 |
| E | 141 hCoV-19/Scotland/EDB2798/2020\|EPI_ISL_487802\|2020- | ···················· | ···················· | ····················· | ···················· | ···················· | ···················· | |
| E | 142 hCoV-19/Scotland/EDB3521/2020\|EPI_ISL_487803\|2020- | ···················· | ···················· | ····················· | ···················· | ···················· | ···················· | |
| E | 143 hCoV-19/Scotland/EDB5412/2020\|EPI_ISL_487804\|2020- | ···················· | ···················A | ····················· | ···················· | ···················· | ···················· | 1 |
| E | 144 hCoV-19/Scotland/EDB2911/2020\|EPI_ISL_487805\|2020- | ···················· | ···················· | ····················· | ···················· | ···················· | ···················· | |
| E | 145 hCoV-19/Scotland/EDB3392/2020\|EPI_ISL_487806\|2020- | ···················· | ···················· | ····················· | ···················· | ···················· | ···················· | |
| E | 146 hCoV-19/Scotland/EDB3375/2020\|EPI_ISL_487807\|2020- | ···················· | ···················A | ····················· | ···················· | ···················· | ···················· | 1 |
| E | 147 hCoV-19/Scotland/EDB5402/2020\|EPI_ISL_487808\|2020- | ···················· | ···················A | ····················· | ···················· | ···················· | ···················· | 1 |
| E | 148 hCoV-19/Scotland/EDB2865/2020\|EPI_ISL_487809\|2020- | ···················· | ···················A | ····················· | ···················· | ···················· | ···················· | 1 |
| E | 149 hCoV-19/Scotland/EDB5858/2020\|EPI_ISL_487810\|2020- | ···················· | ···················· | ····················· | ···················· | ···················· | ···················· | |

| | | | | | | | | |
|---|---|---|---|---|---|---|---|---|
| E | 150 hCoV-19/Scotland/EDB2724/2020\|EPI_ISL_487811\|2020- | ···················· | ···················A | ···················· | ···················· | ···················· | ···················· | 1 |
| E | 151 hCoV-19/Scotland/EDB5577/2020\|EPI_ISL_487812\|2020- | ···················· | ···················A | ···················· | ···················· | ···················· | ···················· | 1 |
| E | 152 hCoV-19/Scotland/EDB3513/2020\|EPI_ISL_487814\|2020- | ···················· | ···················A | ···················· | ···················· | ···················· | ···················· | 1 |
| E | 153 hCoV-19/Scotland/EDB2886/2020\|EPI_ISL_487815\|2020- | ···················· | ···················· | ···················· | ···················· | ···················· | ···················· | |
| E | 154 hCoV-19/Scotland/EDB3111/2020\|EPI_ISL_487816\|2020- | ···················· | ···················· | ···················· | ···················· | ···················· | ···················· | |
| E | 155 hCoV-19/Scotland/EDB5572/2020\|EPI_ISL_487817\|2020- | ···················· | ···················· | ···················· | ···················· | ···················· | ···················· | |
| E | 156 hCoV-19/Scotland/EDB5491/2020\|EPI_ISL_487818\|2020- | ···················· | ···················A | ···················· | ···················· | ···················· | ···················· | 1 |
| E | 157 hCoV-19/Scotland/EDB5431/2020\|EPI_ISL_487819\|2020- | ···················· | ···················A | ···················· | ···················· | ···················· | ···················· | 1 |
| E | 158 hCoV-19/Scotland/EDB5077/2020\|EPI_ISL_487820\|2020- | ···················· | ···················· | ···················· | ···················· | ···················· | ···················· | |
| E | 159 hCoV-19/Scotland/EDB6270/2020\|EPI_ISL_487821\|2020- | ···················· | ···················A | ···················· | ···················· | ···················· | ···················· | 1 |
| E | 160 hCoV-19/Scotland/EDB5629/2020\|EPI_ISL_487822\|2020- | ···················· | ···················· | ···················· | ···················· | ···················· | ···················· | |
| E | 161 hCoV-19/Scotland/EDB3368/2020\|EPI_ISL_487823\|2020- | ···················· | ···················A | ···················· | ···················· | ···················· | ···················· | 1 |
| E | 162 hCoV-19/Scotland/EDB2739/2020\|EPI_ISL_487824\|2020- | ···················· | ···················· | ···················· | ···················· | ···················· | ···················· | |
| E | 163 hCoV-19/Scotland/EDB3425/2020\|EPI_ISL_487825\|2020- | ···················· | ···················A | ···················· | ···················· | ···················· | ···················· | 1 |
| E | 164 hCoV-19/Scotland/EDB3424/2020\|EPI_ISL_487826\|2020- | ···················· | ···················· | ···················· | ···················· | ···················· | ···················· | |
| E | 165 hCoV-19/Scotland/EDB4275/2020\|EPI_ISL_487841\|2020- | ···················· | ···················· | ···················· | ···················· | ···················· | ···················· | |
| E | 166 hCoV-19/Scotland/EDB6413/2020\|EPI_ISL_487842\|2020- | ···················· | ···················· | ···················· | ···················· | ···················· | ···················· | |
| E | 167 hCoV-19/Scotland/EDB5906/2020\|EPI_ISL_487843\|2020- | ···················· | ···················A | ···················· | ···················· | ···················· | ···················· | 1 |
| E | 168 hCoV-19/Scotland/EDB4337/2020\|EPI_ISL_487844\|2020- | ···················· | ···················· | ···················· | ···················· | ···················· | ···················· | |
| E | 169 hCoV-19/Scotland/EDB5048/2020\|EPI_ISL_487845\|2020- | ···················· | ···················· | ···················· | ···················· | ···················· | ···················· | |
| E | 170 hCoV-19/Scotland/EDB2761/2020\|EPI_ISL_487846\|2020- | ···················· | ···················· | ···················· | ···················· | ···················· | ···················· | |
| E | 171 hCoV-19/Scotland/EDB4147/2020\|EPI_ISL_487847\|2020- | ···················· | ···················A | ···················· | ···················· | ···················· | ···················· | 1 |
| E | 172 hCoV-19/Scotland/EDB6407/2020\|EPI_ISL_487848\|2020- | ···················· | ···················· | ···················· | ···················· | ···················· | ···················· | |
| E | 173 hCoV-19/Scotland/EDB5497/2020\|EPI_ISL_487849\|2020- | ···················· | ···················· | ···················· | ···················· | ···················· | ···················· | |
| E | 174 hCoV-19/Scotland/EDB5401/2020\|EPI_ISL_487850\|2020- | ···················· | ···················A | ···················· | ···················· | ···················· | ···················· | 1 |
| E | 175 hCoV-19/Scotland/EDB3052/2020\|EPI_ISL_487851\|2020- | ···················· | ···················· | ···················· | ···················· | ···················· | ···················· | |
| E | 176 hCoV-19/Scotland/EDB3426/2020\|EPI_ISL_487852\|2020- | ···················· | ···················· | ···················· | ···················· | ···················· | ···················· | |
| E | 177 hCoV-19/Scotland/EDB3493/2020\|EPI_ISL_487853\|2020- | ···················· | ·A·················· | ···················· | ···················· | ···················· | ···················· | |
| E | 178 hCoV-19/Scotland/EDB3575/2020\|EPI_ISL_487854\|2020- | ···················· | ···················· | ···················· | ···················· | ···················· | ···················· | |
| E | 179 hCoV-19/Scotland/EDB6420/2020\|EPI_ISL_487855\|2020- | ···················· | ···················· | ···················· | ···················· | ···················· | ···················· | |
| E | 180 hCoV-19/Scotland/EDB5490/2020\|EPI_ISL_487857\|2020- | ···················· | ···················· | ···················· | ···················· | ···················· | ···················· | |
| E | 181 hCoV-19/Scotland/EDB6300/2020\|EPI_ISL_487858\|2020- | ···················· | ···················· | ···················· | ···················· | ···················· | ···················· | |
| E | 182 hCoV-19/Scotland/EDB4330/2020\|EPI_ISL_487859\|2020- | ···················· | ···················· | ···················· | ···················· | ···················· | ···················· | |
| E | 183 hCoV-19/Scotland/EDB5846/2020\|EPI_ISL_487860\|2020- | ···················· | ···················· | ···················· | ···················· | ···················· | ···················· | |
| E | 184 hCoV-19/Scotland/EDB6047/2020\|EPI_ISL_487861\|2020- | ···················· | ···················A | ···················· | ···················· | ···················· | ···················· | 1 |
| E | 185 hCoV-19/Scotland/EDB5800/2020\|EPI_ISL_487862\|2020- | ···················· | ···················· | ···················· | ···················· | ···················· | ···················· | |
| E | 186 hCoV-19/Scotland/EDB3509/2020\|EPI_ISL_487863\|2020- | ···················· | ···················A | ···················· | ···················· | ···················· | ···················· | 1 |
| E | 187 hCoV-19/Scotland/EDB6400/2020\|EPI_ISL_487864\|2020- | ···················· | ···················· | ···················· | ···················· | ···················· | ···················· | |
| E | 188 hCoV-19/Scotland/EDB5651/2020\|EPI_ISL_487865\|2020- | ···················· | ···················· | ···················· | ···················· | ···················· | ···················· | |
| E | 189 hCoV-19/Scotland/EDB6021/2020\|EPI_ISL_487866\|2020- | ···················· | ·A·················· | ···················· | ···················· | ···················· | ···················· | |
| E | 190 hCoV-19/Scotland/EDB3452/2020\|EPI_ISL_487867\|2020- | ···················· | ···················· | ···················· | ···················· | ···················· | ···················· | |
| E | 191 hCoV-19/Scotland/EDB6041/2020\|EPI_ISL_487868\|2020- | ···················· | ···················· | ···················· | ···················· | ···················· | ···················· | |
| E | 192 hCoV-19/Scotland/EDB2714/2020\|EPI_ISL_487869\|2020- | ···················· | ···················· | ···················· | ···················· | ···················· | ···················· | |
| E | 193 hCoV-19/Scotland/EDB3494/2020\|EPI_ISL_487870\|2020- | ···················· | ·A·················· | ···················· | ···················· | ···················· | ···················· | |
| E | 194 hCoV-19/Scotland/EDB3319/2020\|EPI_ISL_487871\|2020- | ···················· | ···················· | ···················· | ···················· | ···················· | ···················· | |
| E | 195 hCoV-19/Scotland/EDB5993/2020\|EPI_ISL_487872\|2020- | ···················· | ···················· | ···················· | ···················· | ···················· | ···················· | |
| E | 196 hCoV-19/Scotland/EDB5501/2020\|EPI_ISL_487873\|2020- | ···················· | ···················· | ···················· | ···················· | ···················· | ···················· | |
| E | 197 hCoV-19/Scotland/EDB3359/2020\|EPI_ISL_487874\|2020- | ···················· | ···················· | ···················· | ···················· | ···················· | ···················· | |
| E | 198 hCoV-19/Scotland/EDB6267/2020\|EPI_ISL_487875\|2020- | ···················· | ·············A······ | ···················· | ···················· | ···················· | ···················· | |
| E | 199 hCoV-19/Scotland/EDB5567/2020\|EPI_ISL_487876\|2020- | ···················· | ···················· | ···················· | ···················· | ···················· | ···················· | |
| E | 200 hCoV-19/Scotland/EDB5990/2020\|EPI_ISL_487877\|2020- | ···················· | ···················· | ···················· | ···················· | ···················· | ···················· | |
| E | 201 hCoV-19/Scotland/EDB4232/2020\|EPI_ISL_487878\|2020- | ···················· | ···················A | ···················· | ···················· | ···················· | ···················· | 1 |
| E | 202 hCoV-19/Scotland/EDB4244/2020\|EPI_ISL_487879\|2020- | ···················· | ···················· | ···················· | ···················· | ···················· | ···················· | |
| E | 203 hCoV-19/Scotland/EDB3546/2020\|EPI_ISL_487880\|2020- | ···················· | ···················· | ···················· | ···················· | ···················· | ···················· | |
| E | 204 hCoV-19/Scotland/EDB5961/2020\|EPI_ISL_487881\|2020- | ···················· | ·A·················· | ···················· | ···················· | ···················· | ···················· | |
| E | 205 hCoV-19/Scotland/EDB3323/2020\|EPI_ISL_487882\|2020- | ···················· | ···················A | ···················· | ···················· | ···················· | ···················· | 1 |
| E | 206 hCoV-19/Scotland/EDB2736/2020\|EPI_ISL_487883\|2020- | ···················· | ···················· | ···················· | ···················· | ···················· | ···················· | |
| E | 207 hCoV-19/Scotland/EDB4717/2020\|EPI_ISL_487884\|2020- | ···················· | ·············A······ | ···················· | ···················· | ···················· | ···················· | |
| E | 208 hCoV-19/Scotland/EDB5931/2020\|EPI_ISL_487885\|2020- | ···················· | ···················· | ···················· | ···················· | ···················· | ···················· | |
| E | 209 hCoV-19/Scotland/EDB3458/2020\|EPI_ISL_487886\|2020- | ···················· | ···················· | ···················· | ···················· | ···················· | ···················· | |
| E | 210 hCoV-19/Scotland/EDB3542/2020\|EPI_ISL_487887\|2020- | ···················· | ···················· | ···················· | ···················· | ···················· | ···················· | |
| E | 211 hCoV-19/Scotland/EDB3413/2020\|EPI_ISL_487888\|2020- | ···················· | ···················· | ···················· | ···················· | ···················· | ···················· | |
| E | 212 hCoV-19/Scotland/EDB3408/2020\|EPI_ISL_487889\|2020- | ···················· | ···················A | ···················· | ···················· | ···················· | ···················· | 1 |
| E | 213 hCoV-19/Scotland/EDB5781/2020\|EPI_ISL_487890\|2020- | ···················· | ···················· | ···················· | ···················· | ···················· | ···················· | |
| E | 214 hCoV-19/Scotland/EDB5807/2020\|EPI_ISL_487891\|2020- | ···················· | ···················A | ···················· | ···················· | ···················· | ···················· | 1 |
| E | 215 hCoV-19/Scotland/EDB5418/2020\|EPI_ISL_487892\|2020- | ···················· | ···················A | ···················· | ···················· | ···················· | ···················· | 1 |
| E | 216 hCoV-19/Scotland/EDB3155/2020\|EPI_ISL_487893\|2020- | ···················· | ·A·················· | ···················· | ···················· | ···················· | ···················· | |
| E | 217 hCoV-19/Scotland/EDB6308/2020\|EPI_ISL_487894\|2020- | ···················· | ···················· | ···················· | ···················· | ···················· | ···················· | |
| E | 218 hCoV-19/Scotland/EDB4309/2020\|EPI_ISL_487895\|2020- | ···················· | ···················· | ···················· | ···················· | ···················· | ···················· | |
| E | 219 hCoV-19/Scotland/EDB3005/2020\|EPI_ISL_487897\|2020- | ···················· | ···················A | ···················· | ···················· | ···················· | ···················· | 1 |
| E | 220 hCoV-19/Scotland/EDB6002/2020\|EPI_ISL_487899\|2020- | ···················· | ···················A | ···················· | ···················· | ···················· | ···················· | 1 |
| E | 221 hCoV-19/Romania/ROSV_3019/2020\|EPI_ISL_491064\|2020 | ···················· | ···················· | ···················· | ···················· | ···················· | ···················· | |
| E | 222 hCoV-19/Romania/ROSV_3169/2020\|EPI_ISL_491065\|2020 | ···················· | ···················· | ···················· | ···················· | ···················· | ···················· | |
| E | 223 hCoV-19/Romania/ROSV_3194/2020\|EPI_ISL_491066\|2020 | ···················· | ···················· | ···················· | ···················· | ···················· | ···················· | |
| E | 224 hCoV-19/Romania/ROSV_3242/2020\|EPI_ISL_491067\|2020 | ···················· | ···················· | ···················· | ···················· | ···················· | ···················· | |
| E | 225 hCoV-19/Romania/ROSV_3376/2020\|EPI_ISL_491069\|2020 | ···················· | ···················· | ···················· | ···················· | ···················· | ···················· | |
| E | 226 hCoV-19/Romania/ROSV_3561/2020\|EPI_ISL_491070\|2020 | ···················· | ···················· | ···················· | ···················· | ···················· | ···················· | |
| E | 227 hCoV-19/Romania/ROSV_3962/2020\|EPI_ISL_491072\|2020 | ···················· | ···················· | ···················· | ···················· | ···················· | ···················· | |
| E | 228 hCoV-19/Romania/ROSV_4271/2020\|EPI_ISL_491073\|2020 | ···················· | ···················· | ···················· | ···················· | ···················· | ···················· | |
| E | 230 hCoV-19/Romania/ROSV_4657/2020\|EPI_ISL_491075\|2020 | ···················· | ···················· | ···················· | ···················· | ···················· | ···················· | |

```
E 231 hCoV-19/Romania/ROSV_4659/2020|EPI_ISL_491076|2020 .................... .................... .................... .................... .................... ....................
E 233 hCoV-19/Romania/ROSV_4714/2020|EPI_ISL_491078|2020 .................... .................... .................... .................... .................... ....................
E 234 hCoV-19/Romania/ROSV_4730/2020|EPI_ISL_491079|2020 .................... .................... .................... .................... .................... ....................
E 235 hCoV-19/Romania/ROSV_4995/2020|EPI_ISL_491080|2020 .................... .................... .................... .................... .................... ....................
E 236 hCoV-19/Romania/ROSV_4996/2020|EPI_ISL_491081|2020 .................... .................... .................... .................... .................... ....................
E 237 hCoV-19/Romania/ROSV_5000/2020|EPI_ISL_491082|2020 .................... .................... .................... .................... .................... ....................
E 239 hCoV-19/Romania/ROSV_5259/2020|EPI_ISL_491084|2020 .................... .................... .................... .................... .................... ....................
E 240 hCoV-19/Romania/ROSV_569/2020|EPI_ISL_491085|2020- .................... .................... .................... .................... .................... ....................
E 241 hCoV-19/USA/SC-DHEC-0001/2020|EPI_ISL_491097|2020- .................... .................... .................... .................... .................... ....................
E 242 hCoV-19/USA/SC-DHEC-0003/2020|EPI_ISL_491098|2020- .................... .................... .................... .................... .................... ....................
E 243 hCoV-19/USA/SC-DHEC-0008/2020|EPI_ISL_491099|2020- .................... .................... .................... .................... .................... ....................
E 244 hCoV-19/England/BRIS-1853F33/2020|EPI_ISL_492395|2 .................... .................... .................... .................... .................... ....................
E 245 hCoV-19/England/BRIS-1854293/2020|EPI_ISL_492396|2 .................... .................... .................... .................... .................... ....................
E 246 hCoV-19/England/BRIS-1854521/2020|EPI_ISL_492397|2 .................... .................... .................... .................... .................... ....................
E 247 hCoV-19/England/BRIS-1853847/2020|EPI_ISL_492398|2 .................... .................... .................... .................... .................... ....................
E 248 hCoV-19/England/BRIS-18541C3/2020|EPI_ISL_492399|2 .................... .................... .................... .................... .................... ....................
E 249 hCoV-19/England/SHEF-CB6F0/2020|EPI_ISL_493702|202 .................... .................... .................... .................... .................... ....................
E 250 hCoV-19/England/SHEF-CB7A2/2020|EPI_ISL_493703|202 .................... .................... .................... .................... .................... ....................
E 251 hCoV-19/England/SHEF-CBFB9/2020|EPI_ISL_493704|202 .................... .................... .................... .................... .................... ....................
E 252 hCoV-19/England/SHEF-CC02E/2020|EPI_ISL_493705|202 .................... .................... .................... .................... .................... ....................
E 253 hCoV-19/England/SHEF-CC53E/2020|EPI_ISL_493707|202 .................... .................... .................... .................... .................... ....................
E 254 hCoV-19/England/SHEF-CD069/2020|EPI_ISL_493709|202 .................... .................... .................... .................... .................... ....................
E 255 hCoV-19/England/SHEF-CD755/2020|EPI_ISL_493712|202 .................... .................... .................... .................... .................... ....................
E 256 hCoV-19/England/SHEF-CDC83/2020|EPI_ISL_493716|202 .................... .................... .................... .................... .................... ....................
E 257 hCoV-19/Uruguay/UY-182/2020|EPI_ISL_480333|2020-04 .................... .................... .................... .................... .................... ....................
E 258 hCoV-19/USA/WI-UW-369/2020|EPI_ISL_480351|2020-05- .................... .................... .................... .................... .................... ....................
E 259 hCoV-19/USA/WI-UW-370/2020|EPI_ISL_480352|2020-05- .................... .................... .................... .................... .................... ....................
E 260 hCoV-19/USA/WI-UW-371/2020|EPI_ISL_480353|2020-05- .................... .................... .................... .................... .................... ....................
E 261 hCoV-19/USA/WI-UW-372/2020|EPI_ISL_480354|2020-05- .................... .................... .................... .................... .................... ....................
E 262 hCoV-19/USA/WI-UW-373/2020|EPI_ISL_480355|2020-05- .................... .................... .................... .................... .................... ....................
E 263 hCoV-19/USA/WI-UW-374/2020|EPI_ISL_480356|2020-05- .................... .................... .................... .................... .................... ....................
E 264 hCoV-19/USA/WI-UW-375/2020|EPI_ISL_480357|2020-05- .................... .................... .................... .................... .................... ....................
E 265 hCoV-19/USA/IL-UW-376/2020|EPI_ISL_480358|2020-05- .................... .................... .................... .................... .................... ....................
E 266 hCoV-19/USA/WI-UW-377/2020|EPI_ISL_480359|2020-05- .................... .................... .................... .................... .................... ....................
E 267 hCoV-19/USA/WI-UW-378/2020|EPI_ISL_480360|2020-05- .................... .................... .................... .................... .................... ....................
E 268 hCoV-19/USA/IL-UW-379/2020|EPI_ISL_480361|2020-05- .................... .................... .................... .................... .................... ....................
E 269 hCoV-19/USA/IL-UW-380/2020|EPI_ISL_480362|2020-05- .................... .................... .................... .................... .................... ....................
E 270 hCoV-19/USA/WI-UW-381/2020|EPI_ISL_480363|2020-05- .................... .................... .................... .................... .................... ....................
E 271 hCoV-19/USA/WI-UW-382/2020|EPI_ISL_480364|2020-05- .................... .................... .................... .................... .................... ....................
E 272 hCoV-19/USA/IL-UW-383/2020|EPI_ISL_480365|2020-05- .................... .................... .................... .................... .................... ....................
E 273 hCoV-19/USA/WI-UW-384/2020|EPI_ISL_480366|2020-05- .................... .................... .................... .................... .................... ....................
E 274 hCoV-19/USA/IL-UW-385/2020|EPI_ISL_480367|2020-05- .................... .................... .................... .................... .................... ....................
E 275 hCoV-19/USA/WI-UW-386/2020|EPI_ISL_480368|2020-05- .................... .................... .................... .................... .................... ....................
E 276 hCoV-19/USA/WI-UW-387/2020|EPI_ISL_480369|2020-05- .................... .................... .................... .................... .................... ....................
E 277 hCoV-19/USA/WI-UW-388/2020|EPI_ISL_480370|2020-05- .................... .................... .................... .................... .................... ....................
E 278 hCoV-19/USA/WI-UW-389/2020|EPI_ISL_480371|2020-05- .................... .................... .................... .................... .................... ....................
E 279 hCoV-19/USA/WI-UW-390/2020|EPI_ISL_480372|2020-05- .................... .................... .................... .................... .................... ....................
E 280 hCoV-19/USA/WI-UW-391/2020|EPI_ISL_480373|2020-05- .................... .................... .................... .................... .................... ....................
E 281 hCoV-19/USA/WI-UW-393/2020|EPI_ISL_480375|2020-05- .................... .................... .................... .................... .................... ....................
E 282 hCoV-19/USA/WI-UW-394/2020|EPI_ISL_480376|2020-05- .................... .................... .................... .................... .................... ....................
E 283 hCoV-19/USA/WI-UW-396/2020|EPI_ISL_480378|2020-03- .................... .................... .................... .................... .................... ....................
E 284 hCoV-19/USA/WI-UW-397/2020|EPI_ISL_480379|2020-03- .................... .................... .................... .................... .................... ....................
E 285 hCoV-19/USA/WI-UW-398/2020|EPI_ISL_480380|2020-03- .................... .................... .................... .................... .................... ....................
E 286 hCoV-19/USA/WI-UW-399/2020|EPI_ISL_480381|2020-03- .................... .................... .................... .................... .................... ....................
E 287 hCoV-19/USA/WI-UW-400/2020|EPI_ISL_480382|2020-03- .................... .................... .................... .................... .................... ....................
E 288 hCoV-19/USA/WI-UW-401/2020|EPI_ISL_480383|2020-03- .................... .................... .................... .................... .................... ....................
E 289 hCoV-19/USA/WI-UW-402/2020|EPI_ISL_480384|2020-03- .................... .................... .................... .................... .................... ....................
E 290 hCoV-19/USA/WI-UW-403/2020|EPI_ISL_480385|2020-03- .................... .................... .................... .................... .................... ....................
E 291 hCoV-19/USA/WI-UW-404/2020|EPI_ISL_480386|2020-03- .................... .................... .................... .................... .................... ....................
E 292 hCoV-19/USA/WI-UW-405/2020|EPI_ISL_480387|2020-03- .................... .................... .................... .................... .................... ....................
E 293 hCoV-19/USA/WI-UW-406/2020|EPI_ISL_480388|2020-03- .................... .................... .................... .................... .................... ....................
E 294 hCoV-19/USA/WI-UW-407/2020|EPI_ISL_480389|2020-03- .................... .................... .................... .................... .................... ....................
E 295 hCoV-19/USA/WI-UW-408/2020|EPI_ISL_480390|2020-03- .................... .................... .................... .................... .................... ....................
E 296 hCoV-19/USA/WI-UW-409/2020|EPI_ISL_480391|2020-03- .................... .................... .................... .................... .................... ....................
E 297 hCoV-19/USA/WI-UW-410/2020|EPI_ISL_480392|2020-03- .................... .................... .................... .................... .................... ....................
E 298 hCoV-19/USA/WI-UW-412/2020|EPI_ISL_480394|2020-04- .................... .................... .................... .................... .................... ....................
E 299 hCoV-19/USA/WI-UW-413/2020|EPI_ISL_480395|2020-04- .................... .................... .................... .................... .................... ....................
E 300 hCoV-19/USA/WI-UW-414/2020|EPI_ISL_480396|2020-04- .................... .................... .................... .................... .................... ....................
E 301 hCoV-19/USA/WI-UW-415/2020|EPI_ISL_480397|2020-04- .................... .................... .................... .................... .................... ....................
E 302 hCoV-19/USA/WI-UW-416/2020|EPI_ISL_480398|2020-04- .................... .................... .................... .................... .................... ....................
E 303 hCoV-19/USA/WI-UW-417/2020|EPI_ISL_480399|2020-04- .................... .................... .................... .................... .................... ....................
E 304 hCoV-19/Finland/26A48S5/2020|EPI_ISL_481620|2020-0 .................... .................... .................... .................... .................... ....................
E 305 hCoV-19/Finland/26A79S5/2020|EPI_ISL_481621|2020-0 .................... .................... .................... .................... .................... ....................
E 306 hCoV-19/Finland/26M7124/2020|EPI_ISL_481622|2020-0 .................... .................... .................... .................... .................... ....................
E 307 hCoV-19/Finland/28A40S6/2020|EPI_ISL_481625|2020-0 .................... .................... .................... .................... .................... ....................
E 308 hCoV-19/Finland/30M83/2020|EPI_ISL_481660|2020-03- .................... .................... .................... .................... .................... ....................
E 309 hCoV-19/Finland/30M88S2/2020|EPI_ISL_481661|2020-0 .................... .................... .................... .................... .................... ....................
E 310 hCoV-19/Finland/30MA7S2/2020|EPI_ISL_481662|2020-0 .................... .................... .................... .................... .................... ....................
E 311 hCoV-19/Finland/30MBS2/2020|EPI_ISL_481663|2020-03 .................... .................... .................... .................... .................... ....................
E 312 hCoV-19/Finland/3A19S9/2020|EPI_ISL_481664|2020-04 .................... .................... .................... .................... .................... ....................
```

| | | | | | | | | | |
|---|---|---|---|---|---|---|---|---|---|
| E | 313 | hCoV-19/Finland/3A21S9/2020\|EPI_ISL_481665\|2020-04 | .................... | .................... | .................... | .................... | .................... | .................... | |
| E | 314 | hCoV-19/Finland/3A26S9/2020\|EPI_ISL_481666\|2020-04 | .................... | .................... | .................... | .................... | .................... | .................... | |
| E | 315 | hCoV-19/Finland/3A47S9/2020\|EPI_ISL_481667\|2020-04 | .................... | .................... | .................... | .................... | .................... | .................... | |
| E | 316 | hCoV-19/Finland/3A56S9/2020\|EPI_ISL_481668\|2020-04 | .................... | .................... | .................... | .................... | .................... | .................... | |
| E | 317 | hCoV-19/Finland/4A33S1/2020\|EPI_ISL_481669\|2020-04 | .................... | .................... | .................... | .................... | .................... | .................... | |
| E | 318 | hCoV-19/Finland/4A60S1/2020\|EPI_ISL_481670\|2020-04 | .................... | .................... | .................... | .................... | .................... | .................... | |
| E | 319 | hCoV-19/Finland/4A70S1/2020\|EPI_ISL_481671\|2020-04 | .................... | .................... | .................... | .................... | .................... | .................... | |
| E | 320 | hCoV-19/Finland/4A75S1/2020\|EPI_ISL_481672\|2020-04 | .................... | .................... | .................... | .................... | .................... | .................... | |
| E | 321 | hCoV-19/Finland/4A9S1/2020\|EPI_ISL_481673\|2020-04- | .................... | .................... | .................... | .................... | .................... | .................... | |
| E | 322 | hCoV-19/Finland/4May21S4/2020\|EPI_ISL_481674\|2020- | .................... | .................... | .................... | .................... | .................... | .................... | |
| E | 323 | hCoV-19/Finland/4May22S4/2020\|EPI_ISL_481675\|2020- | .................... | .................... | .................... | .................... | .................... | .................... | |
| E | 324 | hCoV-19/Finland/4May26S4/2020\|EPI_ISL_481676\|2020- | .................... | .................... | .................... | .................... | .................... | .................... | |
| E | 325 | hCoV-19/Finland/4May27S4/2020\|EPI_ISL_481677\|2020- | .................... | .................... | .................... | .................... | .................... | .................... | |
| E | 326 | hCoV-19/Finland/4May56S4/2020\|EPI_ISL_481678\|2020- | .................... | .................... | .................... | .................... | .................... | .................... | |
| E | 327 | hCoV-19/Finland/4May88S4/2020\|EPI_ISL_481681\|2020- | .................... | .................... | .................... | .................... | .................... | .................... | |
| E | 328 | hCoV-19/Finland/5A10S4A10/2020\|EPI_ISL_481682\|2020 | .................... | .................... | .................... | .................... | .................... | .................... | |
| E | 329 | hCoV-19/Finland/5A31S4A10/2020\|EPI_ISL_481683\|2020 | .................... | .................... | .................... | .................... | .................... | .................... | |
| E | 330 | hCoV-19/Finland/5A34S4A10/2020\|EPI_ISL_481684\|2020 | .................... | .................... | .................... | .................... | .................... | .................... | |
| E | 331 | hCoV-19/Finland/5A40S4A10/2020\|EPI_ISL_481685\|2020 | .................... | .................... | .................... | .................... | .................... | G................... | |
| E | 332 | hCoV-19/Finland/5A44S2/2020\|EPI_ISL_481686\|2020-04 | .................... | .................... | .................... | .................... | .................... | .................... | |
| E | 333 | hCoV-19/Finland/5A49S4A10/2020\|EPI_ISL_481687\|2020 | .................... | .................... | .................... | .................... | .................... | .................... | |
| E | 334 | hCoV-19/Finland/5A4S4A10/2020\|EPI_ISL_481688\|2020- | .................... | .................... | .................... | .................... | .................... | .................... | |
| E | 335 | hCoV-19/Finland/5A55S2/2020\|EPI_ISL_481689\|2020-04 | .................... | .................... | .................... | .................... | .................... | .................... | |
| E | 336 | hCoV-19/Finland/5A57S2/2020\|EPI_ISL_481690\|2020-04 | .................... | .................... | .................... | .................... | .................... | .................... | |
| E | 337 | hCoV-19/Finland/5A57S4A10/2020\|EPI_ISL_481691\|2020 | .................... | .................... | .................... | .................... | .................... | .................... | |
| E | 338 | hCoV-19/Finland/5A71S4A10/2020\|EPI_ISL_481692\|2020 | .................... | .................... | .................... | .................... | .................... | .................... | |
| E | 339 | hCoV-19/Finland/5A81S4A10/2020\|EPI_ISL_481693\|2020 | .................... | .................... | .................... | .................... | .................... | .................... | |
| E | 340 | hCoV-19/Finland/5May13S12/2020\|EPI_ISL_481694\|2020 | .................... | .................... | .................... | .................... | .................... | .................... | |
| E | 341 | hCoV-19/Finland/5May16S2/2020\|EPI_ISL_481695\|2020- | .................... | .................... | .................... | .................... | .................... | .................... | |
| E | 342 | hCoV-19/Finland/5May25S4/2020\|EPI_ISL_481696\|2020- | .................... | .................... | .................... | .................... | .................... | .................... | |
| E | 343 | hCoV-19/Finland/5May26S2/2020\|EPI_ISL_481697\|2020- | .................... | .................... | .................... | .................... | .................... | .................... | |
| E | 344 | hCoV-19/Finland/5May36S12/2020\|EPI_ISL_481698\|2020 | .................... | .................... | .................... | .................... | .................... | .................... | |
| E | 345 | hCoV-19/Finland/5May44S12/2020\|EPI_ISL_481699\|2020 | .................... | .................... | .................... | .................... | .................... | .................... | |
| E | 346 | hCoV-19/USA/MN-MDH-1229/2020\|EPI_ISL_482950\|2020-0 | .................... | .................... | .................... | .................... | .................... | .................... | |
| E | 347 | hCoV-19/USA/MN-MDH-1230/2020\|EPI_ISL_482951\|2020-0 | .................... | .................... | .................... | .................... | .................... | .................... | |
| E | 348 | hCoV-19/USA/MN-MDH-1231/2020\|EPI_ISL_482952\|2020-0 | .................... | .................... | .................... | .................... | .................... | .................... | |
| E | 349 | hCoV-19/USA/MN-MDH-1232/2020\|EPI_ISL_482953\|2020-0 | .................... | .................... | .................... | .................... | .................... | .................... | |
| E | 350 | hCoV-19/USA/MN-MDH-1235/2020\|EPI_ISL_482956\|2020-0 | .................... | .................... | .................... | .................... | .................... | .................... | |
| E | 351 | hCoV-19/USA/MN-MDH-1236/2020\|EPI_ISL_482957\|2020-0 | .................... | .................... | .................... | .................... | .................... | .................... | |
| E | 352 | hCoV-19/USA/MN-MDH-1269/2020\|EPI_ISL_482990\|2020-0 | .................... | .................... | .................... | .................... | .................... | .................... | |
| E | 353 | hCoV-19/USA/MN-MDH-1270/2020\|EPI_ISL_482991\|2020-0 | .................... | .................... | .................... | .................... | .................... | .................... | |
| E | 354 | hCoV-19/USA/MN-MDH-1271/2020\|EPI_ISL_482992\|2020-0 | .................... | .................... | .................... | .................... | .................... | .................... | |
| E | 355 | hCoV-19/USA/MN-MDH-1272/2020\|EPI_ISL_482993\|2020-0 | .................... | .................... | .................... | .................... | .................... | .................... | |
| E | 356 | hCoV-19/USA/MN-MDH-1273/2020\|EPI_ISL_482994\|2020-0 | .................... | .................... | .................... | .................... | .................... | .................... | |
| E | 357 | hCoV-19/USA/MN-MDH-1274/2020\|EPI_ISL_482995\|2020-0 | .................... | .................... | .................... | .................... | .................... | .................... | |
| E | 358 | hCoV-19/USA/MN-MDH-1275/2020\|EPI_ISL_482996\|2020-0 | .................... | .................... | .................... | .................... | .................... | .................... | |
| E | 359 | hCoV-19/USA/MN-MDH-1276/2020\|EPI_ISL_482997\|2020-0 | .................... | .................... | .................... | .................... | .................... | .................... | |
| E | 360 | hCoV-19/USA/MN-MDH-1277/2020\|EPI_ISL_482998\|2020-0 | .................... | .................... | .................... | .................... | .................... | .................... | |
| E | 361 | hCoV-19/USA/MN-MDH-1278/2020\|EPI_ISL_482999\|2020-0 | .................... | .................... | .................... | .................... | .................... | .................... | |
| E | 362 | hCoV-19/Switzerland/ZH-ETHZ-160002/2020\|EPI_ISL_48 | .................... | .................... | .................... | .................... | .................... | .................... | |
| E | 363 | hCoV-19/Switzerland/VS-ETHZ-160006/2020\|EPI_ISL_48 | .................... | .................... | .................... | .................... | .................... | .................... | |
| E | 364 | hCoV-19/Switzerland/VS-ETHZ-160007/2020\|EPI_ISL_48 | .................... | .................... | .................... | .................... | .................... | .................... | |
| E | 365 | hCoV-19/Switzerland/ZH-ETHZ-160008/2020\|EPI_ISL_48 | .................... | .................... | .................... | .................... | .................... | .................... | |
| E | 366 | hCoV-19/Switzerland/BL-ETHZ-160010/2020\|EPI_ISL_48 | .................... | .................... | .................... | .................... | .................... | .................... | |
| E | 367 | hCoV-19/Switzerland/ZH-ETHZ-160015/2020\|EPI_ISL_48 | .................... | .................... | .................... | .................... | ..............Y..... | .................... | |
| E | 368 | hCoV-19/Scotland/EDB5855/2020\|EPI_ISL_487827\|2020- | .................... | .................... | .................... | .................... | .................... | .................... | |
| E | 369 | hCoV-19/Scotland/EDB5634/2020\|EPI_ISL_487828\|2020- | .................... | ...................A | .................... | .................... | .................... | .................... | 1 |
| E | 370 | hCoV-19/Scotland/EDB3391/2020\|EPI_ISL_487829\|2020- | .................... | ...................A | .................... | .................... | .................... | .................... | 1 |
| E | 371 | hCoV-19/Scotland/EDB5924/2020\|EPI_ISL_487830\|2020- | .................... | .................... | .................... | .................... | .................... | .................... | |
| E | 372 | hCoV-19/Scotland/EDB3347/2020\|EPI_ISL_487831\|2020- | .................... | .................... | .................... | .................... | .................... | .................... | |
| E | 373 | hCoV-19/Scotland/EDB1402/2020\|EPI_ISL_487832\|2020- | .................... | .................... | .................... | .................... | .................... | .................... | |
| E | 374 | hCoV-19/Scotland/EDB3563/2020\|EPI_ISL_487833\|2020- | .................... | .A.................. | .................... | .................... | .................... | .................... | |
| E | 375 | hCoV-19/Scotland/EDB3411/2020\|EPI_ISL_487834\|2020- | .................... | .................... | .................... | .................... | .................... | .................... | |
| E | 376 | hCoV-19/Scotland/EDB6426/2020\|EPI_ISL_487835\|2020- | .................... | .................... | .................... | .................... | .................... | .................... | |
| E | 377 | hCoV-19/Scotland/EDB6396/2020\|EPI_ISL_487836\|2020- | .................... | .................... | .................... | .................... | .................... | .................... | |
| E | 378 | hCoV-19/Scotland/EDB5492/2020\|EPI_ISL_487837\|2020- | .................... | .................... | .................... | .................... | .................... | .................... | |
| E | 379 | hCoV-19/Scotland/EDB2963/2020\|EPI_ISL_487838\|2020- | .................... | .................... | .................... | .................... | .................... | .................... | |
| E | 380 | hCoV-19/Scotland/EDB4267/2020\|EPI_ISL_487839\|2020- | .................... | .A.................. | .................... | .................... | .................... | .................... | |
| E | 381 | hCoV-19/Scotland/EDB4058/2020\|EPI_ISL_487840\|2020- | .................... | .................... | .................... | .................... | .................... | .................... | |
| E | 382 | hCoV-19/USA/MN-UW-MAYO71/2020\|EPI_ISL_491000\|2020- | .................... | .................... | .................... | .................... | .................... | .................... | |
| E | 383 | hCoV-19/USA/OR-UW-3406/2020\|EPI_ISL_491001\|2020-03 | .................... | .................... | .................... | .................... | .................... | .................... | |
| E | 384 | hCoV-19/USA/UN-UW-3146/2020\|EPI_ISL_491002\|2020-04 | .................... | .................... | .................... | .................... | .................... | .................... | |
| E | 385 | hCoV-19/USA/UN-UW-3202/2020\|EPI_ISL_491003\|2020-03 | .................... | .................... | .................... | .................... | .................... | .................... | |
| E | 386 | hCoV-19/USA/VT-UW-MAYO49/2020\|EPI_ISL_491004\|2020- | .................... | .................... | .................... | .................... | .................... | .................... | |
| E | 387 | hCoV-19/USA/WA-UW-3158/2020\|EPI_ISL_491006\|2020-03 | .................... | .................... | .................... | .................... | .................... | .................... | |
| E | 388 | hCoV-19/USA/WA-UW-3167/2020\|EPI_ISL_491007\|2020-03 | .................... | .................... | .................... | .................... | .................... | .................... | |
| E | 389 | hCoV-19/USA/WA-UW-3186/2020\|EPI_ISL_491009\|2020-03 | .................... | .................... | .................... | .................... | .................... | .................... | |
| E | 390 | hCoV-19/USA/WA-UW-3188/2020\|EPI_ISL_491010\|2020-03 | .................... | .................... | .................... | .................... | .................... | .................... | |
| E | 391 | hCoV-19/USA/WA-UW-3247/2020\|EPI_ISL_491011\|2020-03 | .................... | .................... | .................... | .................... | .................... | .................... | |
| E | 392 | hCoV-19/USA/WA-UW-3249/2020\|EPI_ISL_491012\|2020-03 | .................... | .................... | .................... | .................... | .................... | .................... | |

| | | | | | | | | |
|---|---|---|---|---|---|---|---|---|
| E | 393 | hCoV-19/USA/WA-UW-3259/2020\|EPI_ISL_491013\|2020-03 | ···················· | ···················· | ····················· | ···················· | ···················· | ···················· |
| E | 394 | hCoV-19/USA/WA-UW-3265/2020\|EPI_ISL_491014\|2020-03 | ···················· | ···················· | ····················· | ···················· | ···················· | ···················· |
| E | 395 | hCoV-19/USA/WA-UW-3336/2020\|EPI_ISL_491016\|2020-03 | ···················· | ···················· | ····················· | ···················· | ···················· | ···················· |
| E | 396 | hCoV-19/USA/WA-UW-3340/2020\|EPI_ISL_491017\|2020-03 | ···················· | ···················· | ····················· | ···················· | ···················· | ···················· |
| E | 397 | hCoV-19/USA/WA-UW-3356/2020\|EPI_ISL_491018\|2020-03 | ···················· | ···················· | ····················· | ···················· | ···················· | ···················· |
| E | 398 | hCoV-19/USA/WA-UW-3365/2020\|EPI_ISL_491019\|2020-03 | ···················· | ···················· | ····················· | ···················· | ···················· | ···················· |
| E | 399 | hCoV-19/USA/WA-UW-3369/2020\|EPI_ISL_491020\|2020-03 | ···················· | ···················· | ····················· | ···················· | ···················· | ···················· |
| E | 400 | hCoV-19/USA/WA-UW-3401/2020\|EPI_ISL_491021\|2020-03 | ···················· | ···················· | ····················· | ···················· | ···················· | ···················· |
| E | 401 | hCoV-19/USA/WA-UW-3413/2020\|EPI_ISL_491022\|2020-03 | ···················· | ···················· | ····················· | ···················· | ···················· | ···················· |
| E | 402 | hCoV-19/USA/WA-UW-3415/2020\|EPI_ISL_491023\|2020-03 | ···················· | ···················· | ····················· | ···················· | ···················· | ···················· |
| E | 403 | hCoV-19/USA/WA-UW-3489/2020\|EPI_ISL_491024\|2020-03 | ···················· | ···················· | ····················· | ···················· | ···················· | ···················· |
| E | 404 | hCoV-19/USA/WA-UW-3490/2020\|EPI_ISL_491025\|2020-03 | ···················· | ···················· | ····················· | ···················· | ···················· | ···················· |
| E | 405 | hCoV-19/USA/WA-UW-3493/2020\|EPI_ISL_491026\|2020-03 | ···················· | ···················· | ····················· | ···················· | ···················· | ···················· |
| E | 406 | hCoV-19/USA/WA-UW-3497/2020\|EPI_ISL_491027\|2020-03 | ···················· | ···················· | ····················· | ···················· | ···················· | ···················· |
| E | 407 | hCoV-19/USA/WA-UW-3500/2020\|EPI_ISL_491028\|2020-03 | ···················· | ···················· | ····················· | ···················· | ···················· | ···················· |
| E | 408 | hCoV-19/USA/WA-UW-3511/2020\|EPI_ISL_491029\|2020-03 | ···················· | ···················· | ····················· | ···················· | ···················· | ···················· |
| E | 409 | hCoV-19/USA/WA-UW-3515/2020\|EPI_ISL_491030\|2020-03 | ···················· | ···················· | ····················· | ···················· | ···················· | ···················· |
| E | 410 | hCoV-19/USA/WA-UW-3521/2020\|EPI_ISL_491031\|2020-03 | ···················· | ···················· | ····················· | ···················· | ···················· | ···················· |
| E | 411 | hCoV-19/USA/WA-UW-3529/2020\|EPI_ISL_491032\|2020-03 | ···················· | ···················· | ····················· | ···················· | ···················· | ···················· |
| E | 412 | hCoV-19/USA/WA-UW-3534/2020\|EPI_ISL_491033\|2020-03 | ···················· | ···················· | ····················· | ···················· | ···················· | ···················· |
| E | 413 | hCoV-19/USA/WA-UW-3539/2020\|EPI_ISL_491034\|2020-03 | ···················· | ···················· | ····················· | ···················· | ···················· | ···················· |
| E | 414 | hCoV-19/Romania/ROSV_10626/2020\|EPI_ISL_491036\|202 | ···················· | ···················· | ····················· | ···················· | ···················· | ···················· |
| E | 415 | hCoV-19/Romania/ROSV_11161/2020\|EPI_ISL_491037\|202 | ···················· | ···················· | ····················· | ···················· | ···················· | ···A················ |
| E | 416 | hCoV-19/Romania/ROSV_11967/2020\|EPI_ISL_491038\|202 | ···················· | ···················· | ····················· | ···················· | ···················· | ···················· |
| E | 420 | hCoV-19/Romania/ROSV_12421/2020\|EPI_ISL_491042\|202 | ···················· | ···················· | ····················· | ···················· | ···················· | ···················· |
| E | 421 | hCoV-19/Romania/ROSV_12442/2020\|EPI_ISL_491043\|202 | ···················· | ···················· | ····················· | ···················· | ···················· | ···················· |
| E | 423 | hCoV-19/Romania/ROSV_12615/2020\|EPI_ISL_491045\|202 | ···················· | ···················· | ····················· | ···················· | ···················· | ···················· |
| E | 424 | hCoV-19/Romania/ROSV_12618/2020\|EPI_ISL_491046\|202 | ···················· | ···················· | ····················· | ···················· | ···················· | ···················· |
| E | 425 | hCoV-19/Romania/ROSV_12619/2020\|EPI_ISL_491047\|202 | ···················· | ···················· | ····················· | ···················· | ···················· | ···················· |
| E | 426 | hCoV-19/Romania/ROSV_12723/2020\|EPI_ISL_491048\|202 | ···················· | ···················· | ····················· | ···················· | ···················· | ···················· |
| E | 427 | hCoV-19/Romania/ROSV_12812/2020\|EPI_ISL_491049\|202 | ···················· | ···················· | ····················· | ···················· | ···················· | ···················· |
| E | 428 | hCoV-19/Romania/ROSV_1296/2020\|EPI_ISL_491050\|2020 | ···················· | ···················· | ····················· | ···················· | ···················· | ···················· |
| E | 429 | hCoV-19/Romania/ROSV_13109/2020\|EPI_ISL_491051\|202 | ···················· | ···················· | ····················· | ···················· | ···················· | ···················· |
| E | 431 | hCoV-19/Romania/ROSV_13429/2020\|EPI_ISL_491053\|202 | ···················· | ···················· | ····················· | ···················· | ···················· | ···················· |
| E | 432 | hCoV-19/Romania/ROSV_1345/2020\|EPI_ISL_491054\|2020 | ···················· | ···················· | ····················· | ···················· | ···················· | ···················· |
| E | 436 | hCoV-19/Romania/ROSV_2492/2020\|EPI_ISL_491058\|2020 | ···················· | ···················· | ····················· | ···················· | ···················· | ···················· |
| E | 438 | hCoV-19/Romania/ROSV_2794/2020\|EPI_ISL_491060\|2020 | ···················· | ···················· | ····················· | ···················· | ···················· | ···················· |
| E | 439 | hCoV-19/Romania/ROSV_2873/2020\|EPI_ISL_491061\|2020 | ···················· | ···················· | ·········T··········· | ···················· | ···················· | ···················· |
| E | 440 | hCoV-19/Romania/ROSV_2874/2020\|EPI_ISL_491062\|2020 | ···················· | ···················· | ····················· | ···················· | ···················· | ···················· |
| E | 441 | hCoV-19/Romania/ROSV_2899/2020\|EPI_ISL_491063\|2020 | ···················· | ···················· | ····················· | ·······C············ | ···················· | ···················· |
| E | 443 | hCoV-19/Romania/ROSV_5849/2020\|EPI_ISL_491087\|2020 | ···················· | ···················· | ····················· | ···················· | ···················· | ···················· |
| E | 444 | hCoV-19/Romania/ROSV_7981/2020\|EPI_ISL_491088\|2020 | ···················· | ···················· | ····················· | ···················· | ···················· | ···················· |
| E | 446 | hCoV-19/Romania/ROSV_8990/2020\|EPI_ISL_491090\|2020 | ···················· | ···················· | ····················· | ···················· | ···················· | ···················· |
| E | 448 | hCoV-19/Czech Republic/NRL_3971/2020\|EPI_ISL_49109 | ···················· | ···················· | ····················· | ···················· | ···················· | ···················· |
| E | 449 | hCoV-19/Czech Republic/NRL_4039/2020\|EPI_ISL_49109 | ···················· | ···················· | ····················· | ···················· | ···················· | ···················· |
| E | 450 | hCoV-19/Czech Republic/NRL_4105/2020\|EPI_ISL_49109 | ···················· | ···················· | ····················· | ···················· | ···················· | ···················· |
| E | 451 | hCoV-19/Czech Republic/NRL_4253/2020\|EPI_ISL_49109 | ···················· | ···················· | ····················· | ···················· | ···················· | ···············A···· |
| E | 453 | hCoV-19/England/BRIS-1852FE9/2020\|EPI_ISL_492313\|2 | ···················· | ···················· | ····················· | ···················· | ···················· | ···················· |
| E | 454 | hCoV-19/England/BRIS-1854178/2020\|EPI_ISL_492314\|2 | ···················· | ···················· | ····················· | ···················· | ···················· | ···················· |
| E | 455 | hCoV-19/England/BRIS-1854512/2020\|EPI_ISL_492315\|2 | ···················· | ···················· | ····················· | ···················· | ···················· | ···················· |
| E | 456 | hCoV-19/England/BRIS-18538B0/2020\|EPI_ISL_492318\|2 | ···················· | ···················· | ····················· | ···················· | ···················· | ···················· |
| E | 457 | hCoV-19/England/BRIS-18543DC/2020\|EPI_ISL_492319\|2 | ···················· | ···················· | ····················· | ···················· | ···················· | ···················· |
| E | 458 | hCoV-19/England/BRIS-18543AF/2020\|EPI_ISL_492320\|2 | ···················· | ···················· | ····················· | ···················· | ···················· | ···················· |
| E | 459 | hCoV-19/England/BRIS-185366B/2020\|EPI_ISL_492321\|2 | ···················· | ···················· | ····················· | ···················· | ···················· | ···················· |
| E | 460 | hCoV-19/England/BRIS-18543BE/2020\|EPI_ISL_492322\|2 | ···················· | ···················· | ····················· | ···················· | ···················· | ···················· |
| E | 461 | hCoV-19/England/BRIS-18544AC/2020\|EPI_ISL_492325\|2 | ···················· | ···················· | ····················· | ···················· | ···················· | ···················· |
| E | 462 | hCoV-19/England/BRIS-1853B99/2020\|EPI_ISL_492329\|2 | ···················· | ···················· | ····················· | ···················· | ···················· | ···················· |
| E | 463 | hCoV-19/England/BRIS-18542B1/2020\|EPI_ISL_492331\|2 | ···················· | ···················· | ····················· | ···················· | ···················· | ···················· |
| E | 464 | hCoV-19/England/BRIS-18545A9/2020\|EPI_ISL_492332\|2 | ···················· | ···················· | ····················· | ···················· | ···················· | ···················· |
| E | 465 | hCoV-19/England/BRIS-18532C1/2020\|EPI_ISL_492334\|2 | ···················· | ···················· | ····················· | ···················· | ···················· | ···················· |
| E | 466 | hCoV-19/England/BRIS-18543CD/2020\|EPI_ISL_492335\|2 | ···················· | ···················· | ····················· | ···················· | ···················· | ···················· |
| E | 467 | hCoV-19/England/BRIS-1854309/2020\|EPI_ISL_492336\|2 | ···················· | ···················· | ····················· | ···················· | ···················· | ···················· |
| E | 468 | hCoV-19/England/BRIS-185380B/2020\|EPI_ISL_492337\|2 | ···················· | ···················· | ····················· | ···················· | ···················· | ···················· |
| E | 469 | hCoV-19/England/BRIS-1853DFD/2020\|EPI_ISL_492338\|2 | ···················· | ···················· | ····················· | ···················· | ···················· | ···················· |
| E | 470 | hCoV-19/England/BRIS-1853D1B/2020\|EPI_ISL_492339\|2 | ···················· | ···················· | ····················· | ···················· | ···················· | ···················· |
| E | 471 | hCoV-19/England/BRIS-18530C7/2020\|EPI_ISL_492341\|2 | ···················· | ···················· | ····················· | ···················· | ···················· | ···················· |
| E | 472 | hCoV-19/England/BRIS-1853908/2020\|EPI_ISL_492344\|2 | ···················· | ···················· | ····················· | ···················· | ···················· | ···················· |
| E | 473 | hCoV-19/England/BRIS-1853A14/2020\|EPI_ISL_492346\|2 | ···················· | ···················· | ····················· | ···················· | ···················· | ···················· |
| E | 474 | hCoV-19/England/BRIS-185349E/2020\|EPI_ISL_492347\|2 | ···················· | ···················· | ····················· | ···················· | ···················· | ···················· |
| E | 475 | hCoV-19/England/BRIS-185421B/2020\|EPI_ISL_492348\|2 | ···················· | ···················· | ····················· | ···················· | ···················· | ···················· |
| E | 476 | hCoV-19/England/BRIS-1853EBE/2020\|EPI_ISL_492349\|2 | ···················· | ···················· | ····················· | ···················· | ···················· | ···················· |
| E | 477 | hCoV-19/England/BRIS-185420C/2020\|EPI_ISL_492350\|2 | ···················· | ···················· | ····················· | ···················· | ···················· | ···················· |
| E | 478 | hCoV-19/England/BRIS-1853DA2/2020\|EPI_ISL_492352\|2 | ···················· | ···················· | ····················· | ···················· | ···················· | ···················· |
| E | 479 | hCoV-19/England/BRIS-185371D/2020\|EPI_ISL_492353\|2 | ···················· | ···················· | ····················· | ···················· | ···················· | ···················· |
| E | 480 | hCoV-19/England/BRIS-1854275/2020\|EPI_ISL_492355\|2 | ···················· | ···················· | ····················· | ···················· | ···················· | ···················· |
| E | 481 | hCoV-19/England/BRIS-1853A7E/2020\|EPI_ISL_492359\|2 | ···················· | ···················· | ····················· | ···················· | ···················· | ···················· |
| E | 482 | hCoV-19/England/BRIS-18538CF/2020\|EPI_ISL_492361\|2 | ···················· | ···················· | ····················· | ···················· | ···················· | ···················· |
| E | 483 | hCoV-19/England/BRIS-1853D39/2020\|EPI_ISL_492363\|2 | ···················· | ···················· | ····················· | ···················· | ···················· | ···················· |
| E | 484 | hCoV-19/England/BRIS-1852FF8/2020\|EPI_ISL_492364\|2 | ···················· | ···················· | ····················· | ···················· | ···················· | ···················· |
| E | 485 | hCoV-19/England/BRIS-18536E3/2020\|EPI_ISL_492366\|2 | ···················· | ···················· | ····················· | ···················· | ···················· | ···················· |

| | | | | | | | | |
|---|---|---|---|---|---|---|---|---|
| E | 486 | hCoV-19/England/BRIS-18540D5/2020\|EPI_ISL_492367\|2 | .................... | .................... | ..................... | .................... | .................... | .................... |
| E | 487 | hCoV-19/England/BRIS-1853D75/2020\|EPI_ISL_492370\|2 | .................... | .................... | ..................... | .................... | .................... | .................... |
| E | 488 | hCoV-19/England/BRIS-18531B5/2020\|EPI_ISL_492371\|2 | .................... | .................... | ..................... | .................... | .................... | .................... |
| E | 489 | hCoV-19/England/BRIS-1853EEB/2020\|EPI_ISL_492372\|2 | .................... | .................... | ..................... | .................... | .................... | .................... |
| E | 490 | hCoV-19/England/BRIS-185411E/2020\|EPI_ISL_492373\|2 | .................... | .................... | ..................... | .................... | .................... | .................... |
| E | 491 | hCoV-19/England/BRIS-18545C7/2020\|EPI_ISL_492377\|2 | .................... | .................... | ..................... | .................... | .................... | .................... |
| E | 492 | hCoV-19/England/BRIS-1854266/2020\|EPI_ISL_492381\|2 | .................... | .................... | ..................... | .................... | .................... | .................... |
| E | 493 | hCoV-19/England/BRIS-1854169/2020\|EPI_ISL_492383\|2 | .................... | .................... | ..................... | .................... | .................... | .................... |
| E | 494 | hCoV-19/England/BRIS-1853E72/2020\|EPI_ISL_492386\|2 | .................... | .................... | ..................... | .................... | .................... | .................... |
| E | 495 | hCoV-19/England/BRIS-1853AC9/2020\|EPI_ISL_492388\|2 | .................... | .................... | ..................... | .................... | .................... | .................... |
| E | 496 | hCoV-19/England/BRIS-1853F24/2020\|EPI_ISL_492390\|2 | .................... | .................... | ..................... | .................... | .................... | .................... |
| E | 497 | hCoV-19/England/BRIS-1853D57/2020\|EPI_ISL_492392\|2 | .................... | .................... | ..................... | .................... | .................... | .................... |
| E | 498 | hCoV-19/England/BRIS-185374A/2020\|EPI_ISL_492393\|2 | .................... | .................... | ..................... | .................... | .................... | .................... |
| E | 499 | hCoV-19/England/PORT-2CF9E2/2020\|EPI_ISL_493614\|20 | .................... | .................... | ..................... | .................... | .................... | .................... |
| E | 500 | hCoV-19/England/PORT-2CFA0D/2020\|EPI_ISL_493615\|20 | .................... | .................... | ..................... | .................... | .................... | .................... |
| E | 501 | hCoV-19/England/PORT-2CFA3A/2020\|EPI_ISL_493617\|20 | .................... | .................... | ..................... | .................... | .................... | .................... |
| E | 502 | hCoV-19/England/SHEF-C17D9/2020\|EPI_ISL_493669\|202 | .................... | .................... | ..................... | .................... | .................... | .................... |
| E | 503 | hCoV-19/England/SHEF-C2C9D/2020\|EPI_ISL_493673\|202 | .................... | .................... | ..................... | .................... | .................... | .................... |
| E | 504 | hCoV-19/England/SHEF-C43A5/2020\|EPI_ISL_493678\|202 | .................... | .................... | ..................... | .................... | .................... | .................... |
| E | 505 | hCoV-19/England/SHEF-C591B/2020\|EPI_ISL_493682\|202 | .................... | .................... | ..................... | .................... | .................... | .................... |
| E | 506 | hCoV-19/England/SHEF-C5CB8/2020\|EPI_ISL_493683\|202 | .................... | .................... | ..................... | .................... | .................... | .................... |
| E | 507 | hCoV-19/England/SHEF-C6051/2020\|EPI_ISL_493684\|202 | .................... | .................... | ..................... | .................... | .................... | .................... |
| E | 508 | hCoV-19/England/SHEF-CA275/2020\|EPI_ISL_493693\|202 | .................... | .................... | ..................... | .................... | .................... | .................... |
| E | 509 | hCoV-19/England/SHEF-CA406/2020\|EPI_ISL_493694\|202 | .................... | .................... | ..................... | .................... | .................... | .................... |
| E | 510 | hCoV-19/England/SHEF-CA9F8/2020\|EPI_ISL_493696\|202 | .................... | .................... | ..................... | .................... | .................... | .................... |
| E | 511 | hCoV-19/England/SHEF-CAD83/2020\|EPI_ISL_493699\|202 | .................... | .................... | ..................... | .................... | .................... | .................... |
| E | 512 | hCoV-19/Canada/LTRI-18/2020\|EPI_ISL_494970\|2020-03 | .................... | .................... | ..................... | .................... | .................... | .................... |
| E | 513 | hCoV-19/Bulgaria/17/2020\|EPI_ISL_480300\|2020-04-14 | .................... | .................... | ..................... | .................... | .................... | .................... |
| E | 514 | hCoV-19/Bulgaria/18/2020\|EPI_ISL_480301\|2020-04-29 | .................... | .................... | ..................... | .................... | .................... | .................... |
| E | 515 | hCoV-19/Bulgaria/21/2020\|EPI_ISL_480302\|2020-05-07 | .................... | .................... | ..................... | .................... | .................... | .................... |
| E | 516 | hCoV-19/Bulgaria/23/2020\|EPI_ISL_480303\|2020-03-30 | .................... | .................... | ..................... | .................... | .................... | .................... |
| E | 517 | hCoV-19/Bulgaria/24/2020\|EPI_ISL_480304\|2020-04-22 | .................... | .................... | ..................... | .................... | .................... | .................... |
| E | 518 | hCoV-19/Bulgaria/27/2020\|EPI_ISL_480305\|2020-05-17 | .................... | .................... | ..................... | .................... | .................... | .................... |
| E | 519 | hCoV-19/Bulgaria/34/2020\|EPI_ISL_480306\|2020-04-25 | .................... | .................... | ..................... | .................... | .................... | .................... |
| E | 520 | hCoV-19/Bulgaria/36/2020\|EPI_ISL_480307\|2020-03-30 | .................... | .................... | ..................... | .................... | .................... | .................... |
| E | 521 | hCoV-19/Bulgaria/37/2020\|EPI_ISL_480308\|2020-05-11 | .................... | .................... | ..................... | .................... | .................... | .................... |
| E | 522 | hCoV-19/Bulgaria/40/2020\|EPI_ISL_480309\|2020-04-19 | .................... | .................... | ..................... | .................... | .................... | .................... |
| E | 523 | hCoV-19/Bulgaria/41/2020\|EPI_ISL_480310\|2020-03-10 | .................... | .................... | ..................... | .................... | .................... | .................... |
| E | 524 | hCoV-19/Finland/26M7974/2020\|EPI_ISL_481623\|2020-0 | .................... | .................... | ..................... | .................... | .................... | .................... |
| E | 525 | hCoV-19/Finland/28A30S6/2020\|EPI_ISL_481624\|2020-0 | .................... | .................... | ..................... | .................... | .................... | .................... |
| E | 526 | hCoV-19/Finland/28A50S11/2020\|EPI_ISL_481626\|2020- | .................... | .................... | ..................... | .................... | .................... | .................... |
| E | 527 | hCoV-19/Finland/28A53S11/2020\|EPI_ISL_481627\|2020- | .................... | .................... | ..................... | .................... | .................... | .................... |
| E | 528 | hCoV-19/Finland/28A54S11/2020\|EPI_ISL_481628\|2020- | .................... | .................... | ..................... | .................... | .................... | .................... |
| E | 529 | hCoV-19/Finland/28A68S17/2020\|EPI_ISL_481629\|2020- | .................... | .................... | ..................... | .................... | .................... | .................... |
| E | 530 | hCoV-19/Finland/28A76S6/2020\|EPI_ISL_481631\|2020-0 | .................... | .................... | ..................... | .................... | .................... | .................... |
| E | 531 | hCoV-19/Finland/28A77S6/2020\|EPI_ISL_481632\|2020-0 | .................... | .................... | ..................... | .................... | .................... | .................... |
| E | 532 | hCoV-19/Finland/28A78S6/2020\|EPI_ISL_481633\|2020-0 | .................... | .................... | ..................... | .................... | .................... | .................... |
| E | 533 | hCoV-19/Finland/28A79S6/2020\|EPI_ISL_481634\|2020-0 | .................... | .................... | ..................... | .................... | .................... | .................... |
| E | 534 | hCoV-19/Finland/28A90S17/2020\|EPI_ISL_481635\|2020- | .................... | .................... | ..................... | .................... | .................... | .................... |
| E | 535 | hCoV-19/Finland/28A92S17/2020\|EPI_ISL_481636\|2020- | .................... | .................... | ..................... | .................... | .................... | .................... |
| E | 536 | hCoV-19/Finland/29A89S/2020\|EPI_ISL_481638\|2020-04 | .................... | .................... | ..................... | .................... | .................... | .................... |
| E | 537 | hCoV-19/Finland/29M11S3/2020\|EPI_ISL_481639\|2020-0 | .................... | .................... | ..................... | .................... | .................... | .................... |
| E | 538 | hCoV-19/Finland/29M14S1/2020\|EPI_ISL_481640\|2020-0 | .................... | .................... | ..................... | .................... | .................... | .................... |
| E | 539 | hCoV-19/Finland/29M15S3/2020\|EPI_ISL_481641\|2020-0 | .................... | .................... | ..................... | .................... | .................... | .................... |
| E | 540 | hCoV-19/Finland/29M17S2/2020\|EPI_ISL_481642\|2020-0 | .................... | .................... | ..................... | .................... | .................... | .................... |
| E | 541 | hCoV-19/Finland/29M18S2/2020\|EPI_ISL_481643\|2020-0 | .................... | .................... | ..................... | .................... | .................... | .................... |
| E | 542 | hCoV-19/Finland/29M36S1/2020\|EPI_ISL_481644\|2020-0 | .................... | .................... | ..................... | .................... | .................... | .................... |
| E | 543 | hCoV-19/Finland/29M50S2/2020\|EPI_ISL_481645\|2020-0 | .................... | .................... | ..................... | .................... | .................... | .................... |
| E | 544 | hCoV-19/Finland/29M60S1/2020\|EPI_ISL_481646\|2020-0 | .................... | .................... | ..................... | .................... | .................... | .................... |
| E | 545 | hCoV-19/Finland/29M60S2/2020\|EPI_ISL_481647\|2020-0 | .................... | .................... | ..................... | .................... | .................... | .................... |
| E | 546 | hCoV-19/Finland/29M77S3/2020\|EPI_ISL_481648\|2020-0 | .................... | .................... | ..................... | .................... | .................... | .................... |
| E | 547 | hCoV-19/Finland/2A92SVIIC/2020\|EPI_ISL_481649\|2020 | .................... | .................... | ..................... | .................... | .................... | .................... |
| E | 548 | hCoV-19/Finland/2A10SVIIC/2020\|EPI_ISL_481650\|2020 | .................... | .A.................. | ..................... | .................... | .................... | .................... |
| E | 549 | hCoV-19/Finland/2A68SVIIIA/2020\|EPI_ISL_481651\|202 | .................... | .................... | ..................... | .................... | .................... | .................... |
| E | 550 | hCoV-19/Finland/2A71SVIIIA/2020\|EPI_ISL_481652\|202 | .................... | .................... | ..................... | .................... | .................... | .................... |
| E | 551 | hCoV-19/Finland/2A72SVIIIA/2020\|EPI_ISL_481653\|202 | .................... | .................... | ..................... | .................... | .................... | .................... |
| E | 552 | hCoV-19/Finland/2A77SVIIIA/2020\|EPI_ISL_481654\|202 | .................... | .................... | ..................... | .................... | .................... | .................... |
| E | 553 | hCoV-19/Finland/2A82SVIIIA/2020\|EPI_ISL_481655\|202 | .................... | .................... | ..................... | .................... | .................... | .................... |
| E | 554 | hCoV-19/Finland/30M6S2/2020\|EPI_ISL_481656\|2020-03 | .................... | .................... | ..................... | .................... | .................... | .................... |
| E | 555 | hCoV-19/Finland/30M75S3/2020\|EPI_ISL_481657\|2020-0 | .................... | .................... | ..................... | .................... | .................... | .................... |
| E | 556 | hCoV-19/Finland/30M77S3/2020\|EPI_ISL_481658\|2020-0 | .................... | .................... | ..................... | .................... | .................... | .................... |
| E | 557 | hCoV-19/Finland/30M80S3/2020\|EPI_ISL_481659\|2020-0 | .................... | .................... | ..................... | .................... | .................... | .................... |
| E | 558 | hCoV-19/England/BRIS-12D7E9/2020\|EPI_ISL_481800\|20 | .................... | .................... | ..................... | .................... | .................... | .................... |
| E | 559 | hCoV-19/England/BRIS-12CF00/2020\|EPI_ISL_481801\|20 | .................... | ....A............... | ..................... | .................... | .................... | .................... |
| E | 560 | hCoV-19/England/BRIS-12CAD2/2020\|EPI_ISL_481803\|20 | .................... | .................... | ..................... | .................... | .................... | .................... |
| E | 561 | hCoV-19/England/BRIS-12D321/2020\|EPI_ISL_481806\|20 | .................... | .................... | ..................... | .................... | .................... | .................... |
| E | 562 | hCoV-19/England/BRIS-12CF4C/2020\|EPI_ISL_481807\|20 | .................... | .................... | ..................... | .................... | .................... | .................... |
| E | 563 | hCoV-19/England/BRIS-12D251/2020\|EPI_ISL_481808\|20 | .................... | .................... | ..................... | .................... | .................... | .................... |
| E | 564 | hCoV-19/England/BRIS-12D664/2020\|EPI_ISL_481810\|20 | .................... | .................... | ..................... | .................... | .................... | .................... |
| E | 565 | hCoV-19/England/BRIS-12D2BB/2020\|EPI_ISL_481811\|20 | .................... | .................... | ..................... | .................... | .................... | .................... |

| | | | | | | | | |
|---|---|---|---|---|---|---|---|---|
| E | 566 | hCoV-19/England/BRIS-12CF1F/2020\|EPI_ISL_481812\|20 | ···················· | ···················· | ····················· | ···················· | ···················· | ···················· |
| E | 567 | hCoV-19/England/BRIS-12DB92/2020\|EPI_ISL_481813\|20 | ···················· | ···················· | ····················· | ···················· | ···················· | ···················· |
| E | 568 | hCoV-19/England/BRIS-12CEE5/2020\|EPI_ISL_481815\|20 | ···················· | ···················· | ····················· | ···················· | ···················· | ···················· |
| E | 569 | hCoV-19/England/BRIS-12CAC3/2020\|EPI_ISL_481817\|20 | ···················· | ···················· | ····················· | ···················· | ···················· | ···················· |
| E | 570 | hCoV-19/England/BRIS-12CBDF/2020\|EPI_ISL_481818\|20 | ···················· | ···················· | ····················· | ···················· | ···················· | ···················· |
| E | 571 | hCoV-19/England/BRIS-12CD51/2020\|EPI_ISL_481819\|20 | ···················· | ···················· | ····················· | ···················· | ···················· | ···················· |
| E | 572 | hCoV-19/England/BRIS-12CB57/2020\|EPI_ISL_481820\|20 | ···················· | ···················· | ····················· | ···················· | ···················· | ···················· |
| E | 573 | hCoV-19/England/BRIS-12DA59/2020\|EPI_ISL_481821\|20 | ···················· | ···················· | ···········T········· | ···················· | ···················· | ···················· |
| E | 574 | hCoV-19/England/BRIS-12CE4F/2020\|EPI_ISL_481822\|20 | ···················· | ···················· | ····················· | ···················· | ···················· | ···················· |
| E | 575 | hCoV-19/England/BRIS-12D9F2/2020\|EPI_ISL_481823\|20 | ···················· | ···················· | ····················· | ···················· | ···················· | ···················· |
| E | 576 | hCoV-19/England/BRIS-12CDE8/2020\|EPI_ISL_481824\|20 | ···················· | ···················· | ····················· | ···················· | ···················· | ···················· |
| E | 577 | hCoV-19/England/BRIS-12D5EF/2020\|EPI_ISL_481825\|20 | ···················· | ···················· | ····················· | ···················· | ···················· | ···················· |
| E | 578 | hCoV-19/England/BRIS-12D594/2020\|EPI_ISL_481826\|20 | ···················· | ···················· | ····················· | ···················· | ···················· | ···················· |
| E | 579 | hCoV-19/England/BRIS-12D88C/2020\|EPI_ISL_481828\|20 | ···················· | ···················· | ····················· | ···················· | ···················· | ···················· |
| E | 580 | hCoV-19/England/BRIS-12D92F/2020\|EPI_ISL_481830\|20 | ···················· | ···················· | ····················· | ···················· | ···················· | ···················· |
| E | 581 | hCoV-19/England/BRIS-12DB0B/2020\|EPI_ISL_481831\|20 | ···················· | ···················· | ····················· | ···················· | ···················· | ···················· |
| E | 582 | hCoV-19/England/BRIS-12CF88/2020\|EPI_ISL_481833\|20 | ···················· | ···················· | ····················· | ···················· | ···················· | ···················· |
| E | 583 | hCoV-19/England/BRIS-12CA4B/2020\|EPI_ISL_481834\|20 | ···················· | ···················· | ····················· | ···················· | ···················· | ···················· |
| E | 584 | hCoV-19/England/BRIS-12D7AD/2020\|EPI_ISL_481836\|20 | ···················· | ···················· | ····················· | ···················· | ···················· | ···················· |
| E | 585 | hCoV-19/England/BRIS-12CB48/2020\|EPI_ISL_481837\|20 | ···················· | ···················· | ····················· | ···················· | ···················· | ···················· |
| E | 586 | hCoV-19/England/BRIS-12CFB5/2020\|EPI_ISL_481839\|20 | ···················· | ···················· | ····················· | ···················· | ···················· | ···················· |
| E | 587 | hCoV-19/England/BRIS-12D3A9/2020\|EPI_ISL_481840\|20 | ···················· | ···················· | ····················· | ···················· | ···················· | ···················· |
| E | 588 | hCoV-19/England/BRIS-12CB1B/2020\|EPI_ISL_481842\|20 | ···················· | ···················· | ····················· | ···················· | ···················· | ···················· |
| E | 589 | hCoV-19/England/BRIS-12D0DF/2020\|EPI_ISL_481843\|20 | ···················· | ···················· | ····················· | ···················· | ···················· | ···················· |
| E | 590 | hCoV-19/England/BRIS-12CF97/2020\|EPI_ISL_481845\|20 | ···················· | ···················· | ····················· | ···················· | ···················· | ···················· |
| E | 591 | hCoV-19/England/BRIS-12D752/2020\|EPI_ISL_481846\|20 | ···················· | ···················· | ····················· | ···················· | ···················· | ···················· |
| E | 592 | hCoV-19/England/BRIS-12CC81/2020\|EPI_ISL_481847\|20 | ···················· | ···················· | ····················· | ···················· | ···················· | ···················· |
| E | 593 | hCoV-19/England/BRIS-12D53A/2020\|EPI_ISL_481848\|20 | ···················· | ···················· | ····················· | ···················· | ···················· | ···················· |
| E | 594 | hCoV-19/England/BRIS-12D707/2020\|EPI_ISL_481850\|20 | ···················· | ···················· | ····················· | ···················· | ···················· | ···················· |
| E | 595 | hCoV-19/England/BRIS-12CC63/2020\|EPI_ISL_481851\|20 | ···················· | ···················· | ····················· | ···················· | ···················· | ···················· |
| E | 596 | hCoV-19/England/BRIS-12D9D4/2020\|EPI_ISL_481852\|20 | ···················· | ···················· | ····················· | ···················· | ···················· | ···················· |
| E | 597 | hCoV-19/England/BRIS-12D048/2020\|EPI_ISL_481853\|20 | ···················· | ···················· | ····················· | ···················· | ···················· | ···················· |
| E | 598 | hCoV-19/England/BRIS-12CC90/2020\|EPI_ISL_481854\|20 | ···················· | ···················· | ····················· | ···················· | ···················· | ···················· |
| E | 599 | hCoV-19/England/BRIS-12D479/2020\|EPI_ISL_481855\|20 | ···················· | ···················· | ····················· | ···················· | ···················· | ···················· |
| E | 600 | hCoV-19/England/BRIS-12D85F/2020\|EPI_ISL_481856\|20 | ···················· | ···················· | ····················· | ···················· | ···················· | ···················· |
| E | 601 | hCoV-19/England/BRIS-12D804/2020\|EPI_ISL_481857\|20 | ···················· | ···················· | ····················· | ···················· | ···················· | ···················· |
| E | 602 | hCoV-19/England/BRIS-12CA69/2020\|EPI_ISL_481858\|20 | ···················· | ···················· | ····················· | ···················· | ···················· | ···················· |
| E | 603 | hCoV-19/England/BRIS-12D233/2020\|EPI_ISL_481859\|20 | ···················· | ···················· | ····················· | ···················· | ···················· | ···················· |
| E | 604 | hCoV-19/USA/MN-MDH-1233/2020\|EPI_ISL_482954\|2020-0 | ···················· | ···················· | ····················· | ···················· | ···················· | ···················· |
| E | 605 | hCoV-19/USA/MN-MDH-1234/2020\|EPI_ISL_482955\|2020-0 | ···················· | ···················· | ····················· | ···················· | ···················· | ···················· |
| E | 606 | hCoV-19/USA/MN-MDH-1237/2020\|EPI_ISL_482958\|2020-0 | ···················· | ···················· | ····················· | ···················· | ···················· | ···················· |
| E | 607 | hCoV-19/USA/MN-MDH-1238/2020\|EPI_ISL_482959\|2020-0 | ···················· | ···················· | ····················· | ···················· | ···················· | ···················· |
| E | 608 | hCoV-19/USA/MN-MDH-1239/2020\|EPI_ISL_482960\|2020-0 | ···················· | ···················· | ····················· | ···················· | ···················· | ···················· |
| E | 609 | hCoV-19/USA/MN-MDH-1240/2020\|EPI_ISL_482961\|2020-0 | ···················· | ···················· | ····················· | ···················· | ···················· | ···················· |
| E | 610 | hCoV-19/USA/MN-MDH-1241/2020\|EPI_ISL_482962\|2020-0 | ···················· | ···················· | ····················· | ···················· | ···················· | ···················· |
| E | 611 | hCoV-19/USA/MN-MDH-1242/2020\|EPI_ISL_482963\|2020-0 | ···················· | ···················· | ····················· | ···················· | ···················· | ···················· |
| E | 612 | hCoV-19/USA/MN-MDH-1243/2020\|EPI_ISL_482964\|2020-0 | ···················· | ···················· | ····················· | ···················· | ···················· | ···················· |
| E | 613 | hCoV-19/USA/MN-MDH-1244/2020\|EPI_ISL_482965\|2020-0 | ···················· | ···················· | ····················· | ···················· | ···················· | ···················· |
| E | 614 | hCoV-19/USA/MN-MDH-1245/2020\|EPI_ISL_482966\|2020-0 | ···················· | ···················· | ····················· | ···················· | ···················· | ···················· |
| E | 615 | hCoV-19/USA/MN-MDH-1246/2020\|EPI_ISL_482967\|2020-0 | ···················· | ···················· | ····················· | ···················· | ···················· | ···················· |
| E | 616 | hCoV-19/USA/MN-MDH-1247/2020\|EPI_ISL_482968\|2020-0 | ···················· | ···················· | ····················· | ···················· | ···················· | ···················· |
| E | 617 | hCoV-19/USA/MN-MDH-1248/2020\|EPI_ISL_482969\|2020-0 | ···················· | ···················· | ····················· | ···················· | ···················· | ···················· |
| E | 618 | hCoV-19/USA/MN-MDH-1249/2020\|EPI_ISL_482970\|2020-0 | ···················· | ···················· | ····················· | ···················· | ···················· | ···················· |
| E | 619 | hCoV-19/USA/MN-MDH-1250/2020\|EPI_ISL_482971\|2020-0 | ···················· | ···················· | ····················· | ···················· | ···················· | ···················· |
| E | 620 | hCoV-19/USA/MN-MDH-1251/2020\|EPI_ISL_482972\|2020-0 | ···················· | ···················· | ····················· | ···················· | ···················· | ···················· |
| E | 621 | hCoV-19/USA/MN-MDH-1252/2020\|EPI_ISL_482973\|2020-0 | ···················· | ···················· | ····················· | ···················· | ···················· | ···················· |
| E | 622 | hCoV-19/USA/MN-MDH-1253/2020\|EPI_ISL_482974\|2020-0 | ···················· | ···················· | ····················· | ···················· | ···················· | ···················· |
| E | 623 | hCoV-19/USA/MN-MDH-1254/2020\|EPI_ISL_482975\|2020-0 | ···················· | ···················· | ····················· | ···················· | ···················· | ···················· |
| E | 624 | hCoV-19/USA/MN-MDH-1255/2020\|EPI_ISL_482976\|2020-0 | ···················· | ···················· | ····················· | ···················· | ···················· | ···················· |
| E | 625 | hCoV-19/USA/MN-MDH-1256/2020\|EPI_ISL_482977\|2020-0 | ···················· | ···················· | ····················· | ···················· | ···················· | ···················· |
| E | 626 | hCoV-19/USA/MN-MDH-1257/2020\|EPI_ISL_482978\|2020-0 | ···················· | ···················· | ····················· | ···················· | ···················· | ···················· |
| E | 627 | hCoV-19/USA/MN-MDH-1258/2020\|EPI_ISL_482979\|2020-0 | ···················· | ···················· | ····················· | ···················· | ···················· | ···················· |
| E | 628 | hCoV-19/USA/MN-MDH-1259/2020\|EPI_ISL_482980\|2020-0 | ···················· | ···················· | ····················· | ···················· | ···················· | ···················· |
| E | 629 | hCoV-19/USA/MN-MDH-1260/2020\|EPI_ISL_482981\|2020-0 | ···················· | ···················· | ····················· | ···················· | ···················· | ···················· |
| E | 630 | hCoV-19/USA/MN-MDH-1261/2020\|EPI_ISL_482982\|2020-0 | ···················· | ···················· | ····················· | ···················· | ···················· | ···················· |
| E | 631 | hCoV-19/USA/MN-MDH-1262/2020\|EPI_ISL_482983\|2020-0 | ···················· | ···················· | ····················· | ···················· | ···················· | ···················· |
| E | 632 | hCoV-19/USA/MN-MDH-1263/2020\|EPI_ISL_482984\|2020-0 | ···················· | ···················· | ····················· | ···················· | ···················· | ···················· |
| E | 633 | hCoV-19/USA/MN-MDH-1264/2020\|EPI_ISL_482985\|2020-0 | ···················· | ···················· | ····················· | ···················· | ···················· | ···················· |
| E | 634 | hCoV-19/USA/MN-MDH-1265/2020\|EPI_ISL_482986\|2020-0 | ···················· | ···················· | ····················· | ···················· | ···················· | ···················· |
| E | 635 | hCoV-19/USA/MN-MDH-1266/2020\|EPI_ISL_482987\|2020-0 | ···················· | ···················· | ····················· | ···················· | ···················· | ···················· |
| E | 636 | hCoV-19/USA/MN-MDH-1267/2020\|EPI_ISL_482988\|2020-0 | ···················· | ···················· | ····················· | ···················· | ···················· | ···················· |
| E | 637 | hCoV-19/USA/MN-MDH-1268/2020\|EPI_ISL_482989\|2020-0 | ···················· | ···················· | ····················· | ···················· | ···················· | ···················· |
| E | 638 | hCoV-19/England/GSTT-265D81F/2020\|EPI_ISL_484019\|2 | ···················· | ···················· | ····················· | ···················· | ···················· | ···················· |
| E | 639 | hCoV-19/England/GSTT-265D82E/2020\|EPI_ISL_484020\|2 | ···················· | ···················· | ····················· | ···················· | ···················· | ···················· |
| E | 640 | hCoV-19/England/GSTT-265D8A6/2020\|EPI_ISL_484028\|2 | ···················· | ···················· | ····················· | ···················· | ···················· | ···················· |
| E | 641 | hCoV-19/England/GSTT-265D90D/2020\|EPI_ISL_484034\|2 | ···················· | ···················· | ····················· | ···················· | ···················· | ···················· |
| E | 642 | hCoV-19/England/GSTT-265D93A/2020\|EPI_ISL_484036\|2 | ···················· | ···················· | ····················· | ···················· | ···················· | ···················· |
| E | 643 | hCoV-19/England/GSTT-265DA46/2020\|EPI_ISL_484053\|2 | ···················· | ···················· | ····················· | ···················· | ···················· | ···················· |
| E | 644 | hCoV-19/England/GSTT-265DA73/2020\|EPI_ISL_484056\|2 | ···················· | ···················· | ····················· | ···················· | ···················· | ···················· |
| E | 645 | hCoV-19/England/GSTT-265DAA0/2020\|EPI_ISL_484059\|2 | ···················· | ···················· | ····················· | ···················· | ···················· | ···················· |

| | | | | | | | | |
|---|---|---|---|---|---|---|---|---|
| E | 646 hCoV-19/England/GSTT-265DAFB/2020\|EPI_ISL_484064\|2 | .................... | .................... | .................... | .................... | .................... | .................... | |
| E | 647 hCoV-19/England/GSTT-265DB52/2020\|EPI_ISL_484070\|2 | .................... | .................... | .................... | .................... | .................... | .................... | |
| E | 648 hCoV-19/England/GSTT-265DB70/2020\|EPI_ISL_484072\|2 | .................... | .................... | .................... | .................... | .................... | .................... | |
| E | 649 hCoV-19/England/GSTT-265DBE9/2020\|EPI_ISL_484079\|2 | .................... | .................... | .................... | .................... | .................... | .................... | |
| E | 650 hCoV-19/England/GSTT-265DC22/2020\|EPI_ISL_484082\|2 | .................... | .................... | .................... | .................... | .................... | .................... | |
| E | 651 hCoV-19/England/GSTT-265DC5F/2020\|EPI_ISL_484085\|2 | .................... | .................... | .................... | .................... | .................... | .................... | |
| E | 652 hCoV-19/England/GSTT-265DC7D/2020\|EPI_ISL_484087\|2 | .................... | .................... | .................... | .................... | .................... | .................... | |
| E | 653 hCoV-19/England/GSTT-265DCB9/2020\|EPI_ISL_484091\|2 | .................... | .................... | .................... | .................... | .................... | .................... | |
| E | 654 hCoV-19/England/GSTT-265DCD7/2020\|EPI_ISL_484093\|2 | .................... | .................... | .................... | .................... | .................... | .................... | |
| E | 655 hCoV-19/England/GSTT-265DD01/2020\|EPI_ISL_484095\|2 | .................... | .................... | .................... | .................... | .................... | .................... | |
| E | 656 hCoV-19/England/GSTT-265DD10/2020\|EPI_ISL_484096\|2 | .................... | .................... | .................... | .................... | .................... | .................... | |
| E | 657 hCoV-19/USA/LA-GBCL-26684489/2020\|EPI_ISL_485300\|2 | .................... | .................... | .................... | .................... | .................... | .................... | |
| E | 658 hCoV-19/USA/LA-GBCL-26684491/2020\|EPI_ISL_485302\|2 | .................... | .................... | .................... | .................... | .................... | .................... | |
| E | 659 hCoV-19/USA/LA-GBCL-26684493/2020\|EPI_ISL_485303\|2 | .................... | .................... | .................... | .................... | .................... | .................... | |
| E | 660 hCoV-19/USA/LA-GBCL-26684496/2020\|EPI_ISL_485304\|2 | .................... | .................... | .................... | .................... | .................... | .................... | |
| E | 661 hCoV-19/USA/LA-GBCL-26684498/2020\|EPI_ISL_485305\|2 | .................... | .................... | .................... | .................... | .................... | .................... | |
| E | 662 hCoV-19/USA/LA-GBCL-26684499/2020\|EPI_ISL_485306\|2 | .................... | .................... | .................... | .................... | .................... | .................... | |
| E | 663 hCoV-19/USA/LA-GBCL-26684505/2020\|EPI_ISL_485310\|2 | .................... | .................... | .................... | .................... | .................... | .................... | |
| E | 664 hCoV-19/USA/LA-GBCL-26684506/2020\|EPI_ISL_485311\|2 | .................... | .................... | .................... | .................... | .................... | .................... | |
| E | 665 hCoV-19/USA/LA-GBCL-26684512/2020\|EPI_ISL_485315\|2 | .................... | .................... | .................... | .................... | .................... | .................... | |
| E | 666 hCoV-19/USA/LA-GBCL-26684513/2020\|EPI_ISL_485316\|2 | .................... | .................... | .................... | .................... | .................... | .................... | |
| E | 667 hCoV-19/USA/LA-GBCL-26684514/2020\|EPI_ISL_485317\|2 | .................... | .................... | .................... | .................... | .................... | .................... | |
| E | 668 hCoV-19/USA/LA-GBCL-26684517/2020\|EPI_ISL_485319\|2 | .................... | .................... | .................... | .................... | .................... | .................... | |
| E | 669 hCoV-19/USA/LA-GBCL-26791694/2020\|EPI_ISL_485320\|2 | .................... | .................... | .................... | .................... | .................... | .................... | |
| E | 670 hCoV-19/USA/LA-GBCL-26791695/2020\|EPI_ISL_485321\|2 | .................... | .................... | .................... | .................... | .................... | .................... | |
| E | 671 hCoV-19/USA/LA-GBCL-26791726/2020\|EPI_ISL_485345\|2 | .................... | .................... | .................... | .................... | .................... | .................... | |
| E | 672 hCoV-19/USA/LA-GBCL-26791734/2020\|EPI_ISL_485351\|2 | .................... | .................... | .................... | .................... | .................... | .................... | |
| E | 673 hCoV-19/USA/LA-GBCL-26791736/2020\|EPI_ISL_485353\|2 | .................... | .................... | .................... | .................... | .................... | .................... | |
| E | 674 hCoV-19/USA/LA-GBCL-26791737/2020\|EPI_ISL_485354\|2 | .................... | .................... | .................... | .................... | .................... | .................... | |
| E | 675 hCoV-19/USA/LA-GBCL-26791739/2020\|EPI_ISL_485356\|2 | .................... | .................... | .................... | .................... | .................... | .................... | |
| E | 676 hCoV-19/USA/LA-GBCL-26791744/2020\|EPI_ISL_485360\|2 | .................... | .................... | .................... | .................... | .................... | .................... | |
| E | 677 hCoV-19/USA/LA-GBCL-26791745/2020\|EPI_ISL_485361\|2 | .................... | .................... | .................... | .................... | .................... | .................... | |
| E | 678 hCoV-19/USA/LA-GBCL-26791749/2020\|EPI_ISL_485364\|2 | .................... | .................... | .................... | .................... | .................... | .................... | |
| E | 679 hCoV-19/USA/LA-GBCL-26791750/2020\|EPI_ISL_485365\|2 | .................... | .................... | .................... | .................... | .................... | .................... | |
| E | 680 hCoV-19/USA/LA-GBCL-26791752/2020\|EPI_ISL_485367\|2 | .................... | .................... | .................... | .................... | .................... | .................... | |
| E | 681 hCoV-19/USA/LA-GBCL-26791753/2020\|EPI_ISL_485368\|2 | .................... | .................... | .................... | .................... | .................... | .................... | |
| E | 682 hCoV-19/USA/LA-GBCL-26791754/2020\|EPI_ISL_485369\|2 | .................... | .................... | .................... | .................... | .................... | .................... | |
| E | 683 hCoV-19/USA/LA-GBCL-26791755/2020\|EPI_ISL_485370\|2 | .................... | .................... | .................... | .................... | .................... | .................... | |
| E | 684 hCoV-19/USA/LA-GBCL-26791756/2020\|EPI_ISL_485371\|2 | .................... | .................... | .................... | .................... | .................... | .................... | |
| E | 685 hCoV-19/USA/LA-GBCL-26791757/2020\|EPI_ISL_485372\|2 | .................... | .................... | .................... | .................... | .................... | .................... | |
| E | 686 hCoV-19/USA/LA-GBCL-26791761/2020\|EPI_ISL_485374\|2 | .................... | .................... | .................... | .................... | .................... | .................... | |
| E | 687 hCoV-19/USA/LA-GBCL-26791762/2020\|EPI_ISL_485375\|2 | .................... | .................... | .................... | .................... | .................... | .................... | |
| E | 688 hCoV-19/USA/LA-GBCL-26791769/2020\|EPI_ISL_485379\|2 | .................... | .................... | .................... | .................... | .................... | .................... | |
| E | 689 hCoV-19/USA/LA-GBCL-26791770/2020\|EPI_ISL_485380\|2 | .................... | .................... | .................... | .................... | ......A............. | .................... | |
| E | 690 hCoV-19/USA/LA-GBCL-26791773/2020\|EPI_ISL_485381\|2 | .................... | .................... | .................... | .................... | .................... | .................... | |
| E | 691 hCoV-19/USA/LA-GBCL-26791774/2020\|EPI_ISL_485382\|2 | .................... | .................... | .................... | .................... | .................... | .................... | |
| E | 692 hCoV-19/USA/LA-GBCL-26791776/2020\|EPI_ISL_485383\|2 | .................... | .................... | .................... | .................... | .................... | .................... | |
| E | 693 hCoV-19/USA/LA-GBCL-26791778/2020\|EPI_ISL_485384\|2 | .................... | .................... | .................... | .................... | .................... | .................... | |
| E | 694 hCoV-19/USA/LA-GBCL-26791779/2020\|EPI_ISL_485385\|2 | .................... | .................... | .................... | .................... | .................... | .................... | |
| E | 695 hCoV-19/USA/LA-GBCL-26791780/2020\|EPI_ISL_485386\|2 | .................... | .................... | .................... | .................... | .................... | .................... | |
| E | 696 hCoV-19/USA/LA-GBCL-26791781/2020\|EPI_ISL_485387\|2 | .................... | .................... | .................... | .................... | .................... | .................... | |
| E | 697 hCoV-19/South Korea/A5/2020\|EPI_ISL_485389\|2020-04 | .................... | .................... | .................... | .................... | .................... | .................... | |
| E | 698 hCoV-19/South Korea/A6/2020\|EPI_ISL_485390\|2020-04 | .................... | .................... | .................... | .................... | .................... | .................... | |
| E | 699 hCoV-19/South Korea/A7/2020\|EPI_ISL_485391\|2020-04 | .................... | .................... | .................... | .................... | .................... | .................... | |
| E | 700 hCoV-19/South Korea/S1/2020\|EPI_ISL_485392\|2020-03 | .................... | .................... | .................... | .................... | .................... | .................... | |
| E | 701 hCoV-19/South Korea/S5/2020\|EPI_ISL_485396\|2020-04 | .................... | .................... | .................... | .................... | .................... | .................... | |
| E | 702 hCoV-19/South Korea/CoV55/2020\|EPI_ISL_485398\|2020 | .................... | .................... | .................... | .................... | .................... | .................... | |
| E | 703 hCoV-19/Poland/IHG_PAS_4_67/2020\|EPI_ISL_485399\|20 | .................... | .................... | .................... | .................... | .................... | .................... | |
| E | 704 hCoV-19/Italy/LOM-ASST-2/2020\|EPI_ISL_486646\|2020- | .................... | .................... | .................... | .................... | .................... | .................... | |
| E | 705 hCoV-19/Italy/LOM-ASST-13/2020\|EPI_ISL_486650\|2020 | .................... | .................... | .................... | .................... | .................... | .................... | |
| E | 706 hCoV-19/Italy/LOM-ASST-14/2020\|EPI_ISL_486651\|2020 | .................... | .................... | .................... | .................... | .................... | .................... | |
| E | 707 hCoV-19/Italy/LOM-ASST-15/2020\|EPI_ISL_486652\|2020 | .................... | .................... | .................... | .................... | .................... | .................... | |
| E | 708 hCoV-19/Italy/LOM-ASST-4/2020\|EPI_ISL_486660\|2020- | .................... | .................... | .................... | .................... | .................... | .................... | |
| E | 709 hCoV-19/Scotland/EDB6305/2020\|EPI_ISL_487962\|2020- | .................... | .............A...... | .................... | .................... | .................... | .................... | |
| E | 710 hCoV-19/Scotland/EDB5429/2020\|EPI_ISL_487963\|2020- | .................... | ...................A | .................... | .................... | .................... | .................... | 1 |
| E | 711 hCoV-19/Scotland/EDB5815/2020\|EPI_ISL_487964\|2020- | .................... | .A.................. | .................... | .................... | .................... | .................... | |
| E | 712 hCoV-19/Scotland/EDB5630/2020\|EPI_ISL_487965\|2020- | .................... | .................... | .................... | .................... | .................... | .................... | |
| E | 713 hCoV-19/Scotland/EDB4829/2020\|EPI_ISL_487966\|2020- | .................... | .................... | .................... | .................... | .................... | .................... | |
| E | 714 hCoV-19/Scotland/EDB3439/2020\|EPI_ISL_487967\|2020- | .................... | .................... | .................... | .................... | .................... | .................... | |
| E | 715 hCoV-19/Scotland/EDB3019/2020\|EPI_ISL_487968\|2020- | .................... | .................... | .................... | .................... | .................... | .................... | |
| E | 716 hCoV-19/Scotland/EDB5658/2020\|EPI_ISL_487969\|2020- | .................... | ...................A | .................... | .................... | .................... | .................... | 1 |
| E | 717 hCoV-19/Scotland/EDB2994/2020\|EPI_ISL_487971\|2020- | .................... | .................... | .................... | .................... | .................... | .................... | |
| E | 718 hCoV-19/Scotland/EDB3543/2020\|EPI_ISL_487972\|2020- | .................... | .................... | .................... | .................... | .................... | .................... | |
| E | 719 hCoV-19/Scotland/EDB5709/2020\|EPI_ISL_487973\|2020- | .................... | .................... | .................... | .................... | .................... | .................... | |
| E | 720 hCoV-19/Scotland/EDB3567/2020\|EPI_ISL_487974\|2020- | .................... | ...................A | .................... | .................... | .................... | .................... | 1 |
| E | 721 hCoV-19/Scotland/EDB3106/2020\|EPI_ISL_487975\|2020- | .................... | ...................A | .................... | .................... | .................... | .................... | 1 |
| E | 722 hCoV-19/Scotland/EDB5794/2020\|EPI_ISL_487976\|2020- | .................... | .................... | .................... | .................... | .................... | .................... | |
| E | 723 hCoV-19/Scotland/EDB6005/2020\|EPI_ISL_487977\|2020- | .................... | ...................A | .................... | .................... | .................... | .................... | 1 |
| E | 724 hCoV-19/Scotland/EDB4322/2020\|EPI_ISL_487979\|2020- | .................... | .................... | .................... | .................... | .................... | .................... | |
| E | 725 hCoV-19/Scotland/EDB3537/2020\|EPI_ISL_487980\|2020- | .................... | ...................A | .................... | .................... | .................... | .................... | 1 |

| | | | | | | | | |
|---|---|---|---|---|---|---|---|---|
| E | 726 hCoV-19/Scotland/EDB6050/2020\|EPI_ISL_487981\|2020- | ···················· | ···················A | ···················· | ···················· | ···················· | ···················· | 1 |
| E | 727 hCoV-19/Scotland/EDB5404/2020\|EPI_ISL_487982\|2020- | ···················· | ···················· | ···················· | ···················· | ···················· | ···················· | |
| E | 728 hCoV-19/Scotland/EDB6301/2020\|EPI_ISL_487983\|2020- | ···················· | ···················· | ···················· | ···················· | ···················· | ···················· | |
| E | 729 hCoV-19/Scotland/EDB5831/2020\|EPI_ISL_487992\|2020- | ···················· | ···················A | ···················· | ···················· | ···················· | ···················· | 1 |
| E | 730 hCoV-19/England/BRIS-1853E63/2020\|EPI_ISL_492300\|2 | ···················· | ···················· | ···················· | ···················· | ···················· | ···················· | |
| E | 731 hCoV-19/England/BRIS-18536C5/2020\|EPI_ISL_492301\|2 | ···················· | ···················· | ···················· | ···················· | ···················· | ···················· | |
| E | 732 hCoV-19/England/BRIS-185322B/2020\|EPI_ISL_492302\|2 | ···················· | ···················· | ···················· | ···················· | ···················· | ···················· | |
| E | 733 hCoV-19/England/BRIS-1854318/2020\|EPI_ISL_492306\|2 | ···················· | ···················· | ···················· | ···················· | ···················· | ···················· | |
| E | 734 hCoV-19/England/BRIS-185372C/2020\|EPI_ISL_492307\|2 | ···················· | ···················· | ···················· | ···················· | ···················· | ···················· | |
| E | 735 hCoV-19/England/BRIS-1853A32/2020\|EPI_ISL_492308\|2 | ···················· | ···················· | ···················· | ···················· | ···················· | ···················· | |
| E | 736 hCoV-19/England/BRIS-185315B/2020\|EPI_ISL_492309\|2 | ···················· | ···················· | ···················· | ···················· | ···················· | ···················· | |
| E | 737 hCoV-19/England/LOND-D7C1D/2020\|EPI_ISL_492500\|202 | ···················· | ···················· | ···················· | ···················· | ···················· | ···················· | |
| E | 738 hCoV-19/England/LOND-D7BE3/2020\|EPI_ISL_492502\|202 | ···················· | ···················· | ···················· | ···················· | ···················· | ···················· | |
| E | 739 hCoV-19/England/LOND-D7BD4/2020\|EPI_ISL_492503\|202 | ···················· | ···················· | ···················· | ···················· | ···················· | ···················· | |
| E | 740 hCoV-19/England/BRIS-130B41/2020\|EPI_ISL_492505\|20 | ···················· | ···················· | ···················· | ···················· | ···················· | ···················· | |
| E | 741 hCoV-19/England/BRIS-130507/2020\|EPI_ISL_492506\|20 | ···················· | ···················· | ···················· | ···················· | ···················· | ···················· | |
| E | 742 hCoV-19/England/BRIS-130CB7/2020\|EPI_ISL_492507\|20 | ···················· | ···················· | ···················· | ···················· | ···················· | ···················· | |
| E | 743 hCoV-19/England/BRIS-130D2D/2020\|EPI_ISL_492508\|20 | ···················· | ·A·················· | ···················· | ···················· | ···················· | ···················· | |
| E | 744 hCoV-19/England/NOTT-112C60/2020\|EPI_ISL_493603\|20 | ···················· | ···················· | ···················· | ···················· | ···················· | ···················· | |
| E | 745 hCoV-19/England/NOTT-112C7F/2020\|EPI_ISL_493604\|20 | ···················· | ···················· | ···················· | ···················· | ···················· | ···················· | |
| E | 746 hCoV-19/England/NOTT-112C9D/2020\|EPI_ISL_493606\|20 | ···················· | ···················· | ···················· | ···················· | ···················· | ···················· | |
| E | 747 hCoV-19/England/NOTT-112CCA/2020\|EPI_ISL_493608\|20 | ···················· | ···················· | ···················· | ···················· | ···················· | ···················· | |
| E | 748 hCoV-19/England/NOTT-112CD9/2020\|EPI_ISL_493609\|20 | ···················· | ···················· | ···················· | ···················· | ···················· | ···················· | |
| E | 749 hCoV-19/England/PORT-2CFA67/2020\|EPI_ISL_493620\|20 | ···················· | ···················· | ···················· | ···················· | ···················· | ···················· | |
| E | 750 hCoV-19/England/PORT-2CFA94/2020\|EPI_ISL_493621\|20 | ···················· | ···················· | ···················· | ···················· | ···················· | ···················· | |
| E | 751 hCoV-19/England/PORT-2CFAB2/2020\|EPI_ISL_493622\|20 | ···················· | ···················· | ···················· | ···················· | ···················· | ···················· | |
| E | 752 hCoV-19/England/PORT-2CFAFE/2020\|EPI_ISL_493624\|20 | ···················· | ···················· | ···················· | ···················· | ···················· | ···················· | |
| E | 753 hCoV-19/England/PORT-2CFB19/2020\|EPI_ISL_493626\|20 | ···················· | ···················· | ···················· | ···················· | ···················· | ···················· | |
| E | 754 hCoV-19/England/PORT-2CFF4A/2020\|EPI_ISL_493628\|20 | ···················· | ···················· | ···················· | ···················· | ···················· | ···················· | |
| E | 755 hCoV-19/England/PORT-2CFF68/2020\|EPI_ISL_493629\|20 | ···················· | ···················· | ···················· | ···················· | ···················· | ···················· | |
| E | 756 hCoV-19/England/PORT-2CFFB3/2020\|EPI_ISL_493633\|20 | ···················· | ···················· | ···················· | ···················· | ···················· | ···················· | |
| E | 757 hCoV-19/England/PORT-2CFFE0/2020\|EPI_ISL_493634\|20 | ···················· | ···················· | ···················· | ···················· | ···················· | ···················· | |
| E | 761 hCoV-19/Scotland/CVR2543/2020\|EPI_ISL_493812\|2020- | ···················· | ···················· | ···················· | ···················· | ···················· | ···················· | |
| E | 762 hCoV-19/Scotland/CVR2547/2020\|EPI_ISL_493814\|2020- | ···················· | ···················A | ···················· | ···················· | ···················· | ···················· | 1 |
| E | 765 hCoV-19/Scotland/CVR2870/2020\|EPI_ISL_493826\|2020- | ···················· | ···················A | ···················· | ···················· | ···················· | ···················· | 1 |
| E | 766 hCoV-19/Scotland/CVR2891/2020\|EPI_ISL_493827\|2020- | ···················· | ·A·················· | ···················· | ···················· | ···················· | ···················· | |
| E | 767 hCoV-19/Scotland/CVR3192/2020\|EPI_ISL_493831\|2020- | ···················· | ···················· | ···················· | ···················· | ···················· | ···················· | |
| E | 769 hCoV-19/Bangladesh/BCSIR-NILMRC-173/2020\|EPI_ISL_4 | ···················· | ···················· | ···················· | ···················· | ···················· | ···················· | |
| E | 770 hCoV-19/England/BRIS-12D190/2020\|EPI_ISL_481781\|20 | ···················· | ·············A······ | ···················· | ···················· | ···················· | ···················· | |
| E | 771 hCoV-19/England/BRIS-12CF6A/2020\|EPI_ISL_481782\|20 | ···················· | ···················· | ···················· | ···················· | ···················· | ···················· | |
| E | 772 hCoV-19/England/BRIS-12D8AA/2020\|EPI_ISL_481783\|20 | ···················· | ···················· | ···················· | ···················· | ···················· | ···················· | |
| E | 773 hCoV-19/England/BRIS-12CB84/2020\|EPI_ISL_481784\|20 | ···················· | ···················· | ···················· | ···················· | ···················· | ···················· | |
| E | 774 hCoV-19/England/BRIS-12D43D/2020\|EPI_ISL_481785\|20 | ···················· | ···················· | ···················· | ···················· | ···················· | ···················· | |
| E | 775 hCoV-19/England/BRIS-12D52B/2020\|EPI_ISL_481786\|20 | ···················· | ···················· | ···················· | ···················· | ···················· | ···················· | |
| E | 776 hCoV-19/England/BRIS-12D6CE/2020\|EPI_ISL_481787\|20 | ···················· | ···················· | ···················· | ···················· | ···················· | ···················· | |
| E | 777 hCoV-19/England/BRIS-12D3B8/2020\|EPI_ISL_481788\|20 | ···················· | ···················· | ···················· | ···················· | ···················· | ···················· | |
| E | 778 hCoV-19/England/BRIS-12D1DC/2020\|EPI_ISL_481789\|20 | ···················· | ···················· | ···················· | ···················· | ···················· | ···················· | |
| E | 779 hCoV-19/England/BRIS-12CC54/2020\|EPI_ISL_481791\|20 | ···················· | ···················· | ···················· | ···················· | ···················· | ···················· | |
| E | 780 hCoV-19/England/BRIS-12D567/2020\|EPI_ISL_481792\|20 | ···················· | ···················· | ···················· | ···················· | ···················· | ···················· | |
| E | 781 hCoV-19/England/BRIS-12CE7C/2020\|EPI_ISL_481793\|20 | ···················· | ···················· | ··T················· | ···················· | ···················· | ···················· | |
| E | 782 hCoV-19/England/BRIS-12CCDC/2020\|EPI_ISL_481794\|20 | ···················· | ···················· | ···················· | ···················· | ···················· | ···················· | |
| E | 783 hCoV-19/England/BRIS-12CCCD/2020\|EPI_ISL_481797\|20 | ···················· | ···················· | ···················· | ···················· | ···················· | ···················· | |
| E | 784 hCoV-19/England/BRIS-12CF79/2020\|EPI_ISL_481799\|20 | ···················· | ···················· | ···················· | ···················· | ···················· | ···················· | |
| E | 785 hCoV-19/England/GSTT-265D703/2020\|EPI_ISL_484002\|2 | ···················· | ···················· | ···················· | ···················· | ···················· | ···················· | |
| E | 786 hCoV-19/England/GSTT-265D730/2020\|EPI_ISL_484005\|2 | ···················· | ···················· | ···················· | ···················· | ···················· | ···················· | |
| E | 787 hCoV-19/England/GSTT-265D76D/2020\|EPI_ISL_484008\|2 | ···················· | ···················· | ···················· | ···················· | ···················· | ···················· | |
| E | 788 hCoV-19/England/GSTT-265D7A9/2020\|EPI_ISL_484012\|2 | ···················· | ···················· | ···················· | ···················· | ···················· | ···················· | |
| E | 789 hCoV-19/USA/LA-GBCL-26791697/2020\|EPI_ISL_485322\|2 | ···················· | ···················· | ···················· | ···················· | ···················· | ···················· | |
| E | 790 hCoV-19/USA/LA-GBCL-26791700/2020\|EPI_ISL_485324\|2 | ···················· | ···················· | ···················· | ···················· | ···················· | ···················· | |
| E | 791 hCoV-19/USA/LA-GBCL-26791704/2020\|EPI_ISL_485327\|2 | ···················· | ···················· | ···················· | ···················· | ···················· | ···················· | |
| E | 792 hCoV-19/USA/LA-GBCL-26791705/2020\|EPI_ISL_485328\|2 | ···················· | ···················· | ···················· | ···················· | ···················· | ···················· | |
| E | 793 hCoV-19/USA/LA-GBCL-26791709/2020\|EPI_ISL_485330\|2 | ···················· | ···················· | ···················· | ···················· | ···················· | ···················· | |
| E | 794 hCoV-19/USA/LA-GBCL-26791711/2020\|EPI_ISL_485331\|2 | ···················· | ···················· | ···················· | ···················· | ···················· | ···················· | |
| E | 795 hCoV-19/USA/LA-GBCL-26791713/2020\|EPI_ISL_485333\|2 | ···················· | ···················· | ···················· | ···················· | ···················· | ···················· | |
| E | 796 hCoV-19/USA/LA-GBCL-26791715/2020\|EPI_ISL_485335\|2 | ···················· | ···················· | ···················· | ···················· | ···················· | ···················· | |
| E | 797 hCoV-19/USA/LA-GBCL-26791719/2020\|EPI_ISL_485338\|2 | ···················· | ···················· | ···················· | ···················· | ···················· | ···················· | |
| E | 798 hCoV-19/USA/LA-GBCL-26791720/2020\|EPI_ISL_485339\|2 | ···················· | ···················· | ···················· | ···················· | ···················· | ···················· | |
| E | 799 hCoV-19/USA/LA-GBCL-26791721/2020\|EPI_ISL_485340\|2 | ···················· | ···················· | ···················· | ···················· | ···················· | ···················· | |
| E | 800 hCoV-19/USA/LA-GBCL-26791723/2020\|EPI_ISL_485342\|2 | ···················· | ···················· | ···················· | ···················· | ···················· | ···················· | |
| E | 801 hCoV-19/USA/LA-GBCL-26791724/2020\|EPI_ISL_485343\|2 | ···················· | ···················· | ···················· | ···················· | ···················· | ···················· | |
| E | 802 hCoV-19/Italy/LOM-ASST-16/2020\|EPI_ISL_486653\|2020 | ···················· | ···················· | ···················· | ···················· | ···················· | ···················· | |
| E | 803 hCoV-19/Italy/LOM-ASST-18/2020\|EPI_ISL_486655\|2020 | ···················· | ···················· | ···················· | ···················· | ···················· | ···················· | |
| E | 804 hCoV-19/Italy/LOM-ASST-20/2020\|EPI_ISL_486657\|2020 | ···················· | ···················· | ···················· | ···················· | ···················· | ···················· | |
| E | 805 hCoV-19/Italy/LOM-ASST-21/2020\|EPI_ISL_486658\|2020 | ···················· | ···················· | ···················· | ···················· | ···················· | ···················· | |
| E | 806 hCoV-19/Italy/LOM-ASST-3/2020\|EPI_ISL_486659\|2020- | ···················· | ···················· | ···················· | ···················· | ···················· | ···················· | |
| E | 807 hCoV-19/Italy/LOM-ASST-6/2020\|EPI_ISL_486662\|2020- | ···················· | ···················· | ···················· | ···················· | ···················· | ···················· | |
| E | 808 hCoV-19/Italy/LOM-ASST-7/2020\|EPI_ISL_486663\|2020- | ···················· | ···················· | ···················· | ···················· | ···················· | ···················· | |
| E | 809 hCoV-19/Italy/LOM-ASST-8/2020\|EPI_ISL_486664\|2020- | ···················· | ···················· | ···················· | ···················· | ···················· | ···················· | |
| E | 810 hCoV-19/Italy/LOM-AMVRC-9/2020\|EPI_ISL_486665\|2020 | ···················· | ···················· | ···················· | ···················· | ···················· | ···················· | |
| E | 811 hCoV-19/India/KA-InStem-NCBS-0094/2020\|EPI_ISL_486 | ···················· | ···················· | ···················· | ···················· | ···················· | ···················· | |

| | | | | | | | | |
|---|---|---|---|---|---|---|---|---|
| E | 812 hCoV-19/India/KA-InStem-NCBS-0095/2020\|EPI_ISL_486 | ···················· | ···················· | ···················· | ···················· | ···················· | ···················· | |
| E | 813 hCoV-19/Scotland/EDB5055/2020\|EPI_ISL_487900\|2020- | ···················· | ···················· | ···················· | ···················· | ···················· | ···················· | |
| E | 814 hCoV-19/Scotland/EDB3561/2020\|EPI_ISL_487901\|2020- | ···················· | ···················A | ···················· | ···················· | ···················· | ···················· | 1 |
| E | 815 hCoV-19/Scotland/EDB6403/2020\|EPI_ISL_487902\|2020- | ···················· | ·A·················· | ···················· | ···················· | ···················· | ···················· | |
| E | 816 hCoV-19/Scotland/EDB5943/2020\|EPI_ISL_487903\|2020- | ···················· | ···················· | ···················· | ···················· | ···················· | ···················· | |
| E | 817 hCoV-19/Scotland/EDB5939/2020\|EPI_ISL_487904\|2020- | ···················· | ···················A | ···················· | ···················· | ···················· | ···················· | 1 |
| E | 818 hCoV-19/Scotland/EDB5912/2020\|EPI_ISL_487905\|2020- | ···················· | ···················A | ···················· | ···················· | ···················· | ···················· | 1 |
| E | 819 hCoV-19/Scotland/EDB5811/2020\|EPI_ISL_487906\|2020- | ···················· | ···················· | ···················· | ···················· | ···················· | ···················· | |
| E | 820 hCoV-19/Scotland/EDB3324/2020\|EPI_ISL_487907\|2020- | ···················· | ···················A | ···················· | ···················· | ···················· | ···················· | 1 |
| E | 821 hCoV-19/Scotland/EDB6414/2020\|EPI_ISL_487908\|2020- | ···················· | ···················· | ···················· | ···················· | ···················· | ···················· | |
| E | 822 hCoV-19/Scotland/EDB3556/2020\|EPI_ISL_487909\|2020- | ···················· | ···················· | ···················· | ···················· | ···················· | ···················· | |
| E | 823 hCoV-19/Scotland/EDB5522/2020\|EPI_ISL_487910\|2020- | ···················· | ···················· | ···················· | ···················· | ···················· | ···················· | |
| E | 824 hCoV-19/Scotland/EDB3315/2020\|EPI_ISL_487911\|2020- | ···················· | ···················A | ···················· | ···················· | ···················· | ···················· | 1 |
| E | 825 hCoV-19/Scotland/EDB5726/2020\|EPI_ISL_487912\|2020- | ···················· | ···················· | ···················· | ···················· | ···················· | ···················· | |
| E | 826 hCoV-19/Scotland/EDB5996/2020\|EPI_ISL_487913\|2020- | ···················· | ···················· | ···················· | ···················· | ···················· | ···················· | |
| E | 827 hCoV-19/Scotland/EDB6275/2020\|EPI_ISL_487914\|2020- | ···················· | ···················· | ···················· | ···················· | ···················· | ···················· | |
| E | 828 hCoV-19/Scotland/EDB3423/2020\|EPI_ISL_487915\|2020- | ···················· | ···················A | ···················· | ···················· | ···················· | ···················· | 1 |
| E | 829 hCoV-19/Scotland/EDB3365/2020\|EPI_ISL_487916\|2020- | ···················· | ···················· | ···················· | ···················· | ···················· | ···················· | |
| E | 830 hCoV-19/Scotland/EDB5525/2020\|EPI_ISL_487917\|2020- | ···················· | ···················· | ···················· | ···················· | ···················· | ···················· | |
| E | 831 hCoV-19/Scotland/EDB2728/2020\|EPI_ISL_487918\|2020- | ···················· | ···················· | ···················· | ···················· | ···················· | ···················· | |
| E | 832 hCoV-19/Scotland/EDB5632/2020\|EPI_ISL_487919\|2020- | ···················· | ···················A | ···················· | ···················· | ···················· | ···················· | 1 |
| E | 833 hCoV-19/Scotland/EDB5946/2020\|EPI_ISL_487920\|2020- | ···················· | ···················· | ···················· | ···················· | ···················· | ···················· | |
| E | 834 hCoV-19/Scotland/EDB5500/2020\|EPI_ISL_487921\|2020- | ···················· | ···················· | ···················· | ···················· | ···················· | ···················· | |
| E | 835 hCoV-19/Scotland/EDB3087/2020\|EPI_ISL_487922\|2020- | ···················· | ···················A | ···················· | ···················· | ···················· | ···················· | 1 |
| E | 836 hCoV-19/Scotland/EDB3378/2020\|EPI_ISL_487923\|2020- | ···················· | ···················· | ···················· | ···················· | ···················· | ···················· | |
| E | 837 hCoV-19/Scotland/EDB3318/2020\|EPI_ISL_487924\|2020- | ···················· | ···················A | ···················· | ···················· | ···················· | ···················· | 1 |
| E | 838 hCoV-19/Scotland/EDB5969/2020\|EPI_ISL_487926\|2020- | ···················· | ···················A | ···················· | ···················· | ···················· | ···················· | 1 |
| E | 839 hCoV-19/Scotland/EDB6271/2020\|EPI_ISL_487927\|2020- | ···················· | ···················· | ···················· | ···················· | ···················· | ···················· | |
| E | 840 hCoV-19/Scotland/EDB3341/2020\|EPI_ISL_487928\|2020- | ···················· | ·A·················· | ···················· | ···················· | ···················· | ···················· | |
| E | 841 hCoV-19/Scotland/EDB3547/2020\|EPI_ISL_487929\|2020- | ···················· | ···················· | ···················· | ···················· | ···················· | ···················· | |
| E | 842 hCoV-19/Scotland/EDB3349/2020\|EPI_ISL_487930\|2020- | ···················· | ·A·················· | ···················· | ···················· | ···················· | ···················· | |
| E | 843 hCoV-19/Scotland/EDB5871/2020\|EPI_ISL_487931\|2020- | ···················· | ···················A | ···················· | ···················· | ···················· | ···················· | 1 |
| E | 844 hCoV-19/Scotland/EDB5519/2020\|EPI_ISL_487932\|2020- | ···················· | ···················· | ···················· | ···················· | ···················· | ···················· | |
| E | 845 hCoV-19/Scotland/EDB2699/2020\|EPI_ISL_487933\|2020- | ···················· | ···················· | ···················· | ···················· | ···················· | ···················· | |
| E | 846 hCoV-19/Scotland/EDB2907/2020\|EPI_ISL_487934\|2020- | ···················· | ···················· | ···················· | ···················· | ···················· | ···················· | |
| E | 847 hCoV-19/Scotland/EDB3418/2020\|EPI_ISL_487935\|2020- | ···················· | ···················· | ···················· | ···················· | ···················· | ···················· | |
| E | 848 hCoV-19/Scotland/EDB3422/2020\|EPI_ISL_487936\|2020- | ···················· | ···················A | ···················· | ···················· | ···················· | ···················· | 1 |
| E | 849 hCoV-19/Scotland/EDB3524/2020\|EPI_ISL_487937\|2020- | ···················· | ···················A | ···················· | ···················· | ···················· | ···················· | 1 |
| E | 850 hCoV-19/Scotland/EDB6433/2020\|EPI_ISL_487938\|2020- | ···················· | ···················· | ···················· | ···················· | ···················· | ···················· | |
| E | 851 hCoV-19/Scotland/EDB5971/2020\|EPI_ISL_487939\|2020- | ···················· | ···················A | ···················· | ···················· | ···················· | ···················· | 1 |
| E | 852 hCoV-19/Scotland/EDB2689/2020\|EPI_ISL_487940\|2020- | ···················· | ···················· | ···················· | ···················· | ···················· | ···················· | |
| E | 853 hCoV-19/Scotland/EDB6430/2020\|EPI_ISL_487941\|2020- | ···················· | ···················· | ···················· | ···················· | ···················· | ···················· | |
| E | 854 hCoV-19/Scotland/EDB3564/2020\|EPI_ISL_487942\|2020- | ···················· | ···················A | ···················· | ···················· | ···················· | ···················· | 1 |
| E | 855 hCoV-19/Scotland/EDB3533/2020\|EPI_ISL_487943\|2020- | ···················· | ···················A | ···················· | ···················· | ···················· | ···················· | 1 |
| E | 856 hCoV-19/Scotland/EDB6431/2020\|EPI_ISL_487944\|2020- | ···················· | ···················· | ···················· | ···················· | ···················· | ···················· | |
| E | 857 hCoV-19/Scotland/EDB5887/2020\|EPI_ISL_487945\|2020- | ···················· | ···················· | ···················· | ···················· | ···················· | ···················· | |
| E | 858 hCoV-19/Scotland/EDB5841/2020\|EPI_ISL_487947\|2020- | ···················· | ···················· | ···················· | ···················· | ···················· | ···················· | |
| E | 859 hCoV-19/Scotland/EDB5937/2020\|EPI_ISL_487948\|2020- | ···················· | ···················A | ···················· | ···················· | ···················· | ···················· | 1 |
| E | 860 hCoV-19/Scotland/EDB3314/2020\|EPI_ISL_487949\|2020- | ···················· | ···················· | ···················· | ···················· | ···················· | ···················· | |
| E | 861 hCoV-19/Scotland/EDB3525/2020\|EPI_ISL_487950\|2020- | ···················· | ···················A | ···················· | ···················· | ···················· | ···················· | 1 |
| E | 862 hCoV-19/Scotland/EDB5986/2020\|EPI_ISL_487951\|2020- | ···················· | ···················· | ···················· | ···················· | ···················· | ···················· | |
| E | 863 hCoV-19/Scotland/EDB3380/2020\|EPI_ISL_487952\|2020- | ···················· | ···················· | ···················· | ···················· | ···················· | ···················· | |
| E | 864 hCoV-19/Scotland/EDB6429/2020\|EPI_ISL_487953\|2020- | ···················· | ···················· | ···················· | ···················· | ···················· | ···················· | |
| E | 865 hCoV-19/Scotland/EDB5529/2020\|EPI_ISL_487954\|2020- | ···················· | ···················A | ···················· | ···················· | ···················· | ···················· | 1 |
| E | 866 hCoV-19/Scotland/EDB4251/2020\|EPI_ISL_487956\|2020- | ···················· | ···················A | ···················· | ···················· | ···················· | ···················· | 1 |
| E | 867 hCoV-19/Scotland/EDB5583/2020\|EPI_ISL_487957\|2020- | ···················· | ···················· | ···················· | ···················· | ···················· | ···················· | |
| E | 868 hCoV-19/Scotland/EDB1403/2020\|EPI_ISL_487958\|2020- | ···················· | ···················· | ···················· | ···················· | ···················· | ···················· | |
| E | 869 hCoV-19/Scotland/EDB3343/2020\|EPI_ISL_487959\|2020- | ···················· | ·A·················· | ···················· | ···················· | ···················· | ···················· | |
| E | 870 hCoV-19/Scotland/EDB5425/2020\|EPI_ISL_487960\|2020- | ···················· | ···················A | ···················· | ···················· | ···················· | ···················· | 1 |
| E | 871 hCoV-19/Scotland/EDB3541/2020\|EPI_ISL_487961\|2020- | ···················· | ···················· | ···················· | ···················· | ···················· | ···················· | |
| E | 872 hCoV-19/Scotland/EDB4263/2020\|EPI_ISL_487984\|2020- | ···················· | ···················A | ···················· | ···················· | ···················· | ···················· | 1 |
| E | 873 hCoV-19/Scotland/EDB5717/2020\|EPI_ISL_487985\|2020- | ···················· | ···················· | ···················· | ···················· | ···················· | ···················· | |
| E | 874 hCoV-19/Scotland/EDB5782/2020\|EPI_ISL_487986\|2020- | ···················· | ···················A | ···················· | ···················· | ···················· | ···················· | 1 |
| E | 875 hCoV-19/Scotland/EDB6281/2020\|EPI_ISL_487987\|2020- | ···················· | ···················· | ···················· | ···················· | ···················· | ···················· | |
| E | 876 hCoV-19/Scotland/EDB3421/2020\|EPI_ISL_487988\|2020- | ···················· | ···················A | ···················· | ···················· | ···················· | ···················· | 1 |
| E | 877 hCoV-19/Scotland/EDB6029/2020\|EPI_ISL_487989\|2020- | ···················· | ···················A | ···················· | ···················· | ···················· | ···················· | 1 |
| E | 878 hCoV-19/Scotland/EDB5928/2020\|EPI_ISL_487990\|2020- | ···················· | ···················· | ···················· | ···················· | ···················· | ···················· | |
| E | 879 hCoV-19/Scotland/EDB2848/2020\|EPI_ISL_487991\|2020- | ···················· | ···················· | ···················· | ···················· | ···················· | ···················· | |
| E | 880 hCoV-19/Scotland/EDB5917/2020\|EPI_ISL_487994\|2020- | ···················· | ·A·················· | ···················· | ···················· | ···················· | ···················· | |
| E | 881 hCoV-19/Scotland/EDB3344/2020\|EPI_ISL_487996\|2020- | ···················· | ···················· | ···················· | ···················· | ···················· | ···················· | |
| E | 882 hCoV-19/Scotland/EDB5494/2020\|EPI_ISL_487997\|2020- | ···················· | ···················· | ···················· | ···················· | ···················· | ···················· | |
| E | 883 hCoV-19/Scotland/EDB3528/2020\|EPI_ISL_487998\|2020- | ···················· | ···················A | ···················· | ···················· | ···················· | ···················· | 1 |
| E | 884 hCoV-19/Scotland/EDB3410/2020\|EPI_ISL_487999\|2020- | ···················· | ·A·················· | ···················· | ···················· | ···················· | ···················· | |
| E | 885 hCoV-19/USA/SC-DHEC-0046/2020\|EPI_ISL_491101\|2020- | ···················· | ···················· | ···················· | ···················· | ···················· | ···················· | |
| E | 886 hCoV-19/USA/SC-DHEC-0048/2020\|EPI_ISL_491102\|2020- | ···················· | ···················· | ···················· | ···················· | ···················· | ···················· | |
| E | 887 hCoV-19/USA/SC-DHEC-0050/2020\|EPI_ISL_491103\|2020- | ···················· | ···················· | ···················· | ···················· | ···················· | ···················· | |
| E | 888 hCoV-19/USA/SC-DHEC-0055/2020\|EPI_ISL_491104\|2020- | ···················· | ···················· | ···················· | ···················· | ···················· | ···················· | |
| E | 889 hCoV-19/USA/SC-DHEC-0076/2020\|EPI_ISL_491105\|2020- | ···················· | ··············C····· | ···················· | ···················· | ···················· | ···················· | |
| E | 890 hCoV-19/USA/SC-DHEC-0077/2020\|EPI_ISL_491106\|2020- | ···················· | ··············C····· | ···················· | ···················· | ···················· | ···················· | |
| E | 891 hCoV-19/USA/SC-DHEC-0085/2020\|EPI_ISL_491107\|2020- | ···················· | ···················· | ···················· | ···················· | ···················· | ···················· | |

| | | | | | | | | |
|---|---|---|---|---|---|---|---|---|
| E | 892 hCoV-19/USA/SC-DHEC-0143/2020\|EPI_ISL_491110\|2020- | ···················· | ·········A·········· | ···················· | ···················· | ···················· | ···················· | |
| E | 893 hCoV-19/India/UN-17074/2020\|EPI_ISL_491114\|2020 | ···················· | ···················· | ···················· | ···················· | ····N····N·········· | ···N············N··· | |
| E | 894 hCoV-19/Germany/NI-HCRI-BrZK-1/2020\|EPI_ISL_491115 | ···················· | ···················· | ···················· | ···················· | ···················· | ···················· | |
| E | 895 hCoV-19/Oman/RESP-20-6020/2020\|EPI_ISL_491116\|2020 | ···················· | ···················· | ···················· | ···················· | ···················· | ···················· | |
| E | 896 hCoV-19/Czech Republic/NRL_5201/2020\|EPI_ISL_49111 | ···················· | ···················· | ···················· | ···················· | ···················· | ···················· | |
| E | 897 hCoV-19/Czech Republic/NRL_5752/2020\|EPI_ISL_49111 | ···················· | ···················· | ···················· | ···················· | ···················· | ···············A···· | |
| E | 898 hCoV-19/Czech Republic/NRL_5815/2020\|EPI_ISL_49111 | ···················· | ···················· | ···················· | ···················· | ···················· | ···················· | |
| E | 899 hCoV-19/Oman/8556/2020\|EPI_ISL_491121\|2020-04-02 | ···················· | ···················· | ···················· | ···················· | ···················· | ···················· | |
| E | 900 hCoV-19/Oman/C-9311/2020\|EPI_ISL_491122\|2020-04-28 | ···················· | ···················· | ···················· | ···················· | ···················· | ···················· | |
| E | 901 hCoV-19/Oman/17309/2020\|EPI_ISL_491123\|2020-04-13 | ···················· | ···················· | ···················· | ···················· | ···················· | ···················· | |
| E | 902 hCoV-19/Oman/C-8871/2020\|EPI_ISL_491124\|2020-04-28 | ···················· | ···················· | ···················· | ···················· | ···················· | ···················· | |
| E | 903 hCoV-19/Oman/C-6175/2020\|EPI_ISL_491125\|2020-04-25 | ···················· | ···················· | ···················· | ···················· | ···················· | ···················· | |
| E | 904 hCoV-19/Oman/C-6324/2020\|EPI_ISL_491126\|2020-04-25 | ···················· | ···················· | ···················· | ···················· | ···················· | ···················· | |
| E | 905 hCoV-19/Oman/C-6583/2020\|EPI_ISL_491127\|2020-04-26 | ···················· | ···················· | ···················· | ···················· | ···················· | ···················· | |
| E | 906 hCoV-19/Oman/8600/2020\|EPI_ISL_491128\|2020-04-03 | ···················· | ···················· | ···················· | ···················· | ···················· | ···················· | |
| E | 907 hCoV-19/Oman/B-2110/2020\|EPI_ISL_491130\|2020-04-21 | ···················· | ···················· | ···················· | ···················· | ···················· | ···················· | |
| E | 908 hCoV-19/Oman/B-3123/2020\|EPI_ISL_491131\|2020-04-25 | ···················· | ···················· | ···················· | ···················· | ···················· | ···················· | |
| E | 909 hCoV-19/Oman/C-6584/2020\|EPI_ISL_491132\|2020-04-26 | ···················· | ···················· | ···················· | ···················· | ···················· | ···················· | |
| E | 910 hCoV-19/Oman/C-9263/2020\|EPI_ISL_491133\|2020-04-28 | ···················· | ···················· | ···················· | ···················· | ···················· | ···················· | |
| E | 911 hCoV-19/Oman/6991/2020\|EPI_ISL_491134\|2020-03-30 | ···················· | ···················· | ···················· | ···················· | ···················· | ···················· | |
| E | 912 hCoV-19/Oman/8290/2020\|EPI_ISL_491135\|2020-04-02 | ···················· | ···················· | ···················· | ···················· | ···················· | ···················· | |
| E | 913 hCoV-19/Oman/C-7102/2020\|EPI_ISL_491136\|2020-04-27 | ···················· | ···················· | ···················· | ···················· | ···················· | ···················· | |
| E | 914 hCoV-19/Oman/20631/2020\|EPI_ISL_491137\|2020-04-15 | ···················· | ···················· | ···················· | ···················· | ···················· | ···················· | |
| E | 915 hCoV-19/Oman/C-22988/2020\|EPI_ISL_491138\|2020-05-1 | ···················· | ···················· | ···················· | ···················· | ···················· | ···················· | |
| E | 916 hCoV-19/Oman/24647/2020\|EPI_ISL_491139\|2020-04-18 | ···················· | ···················· | ···················· | ···················· | ···················· | ···················· | |
| E | 917 hCoV-19/Oman/C-5174/2020\|EPI_ISL_491140\|2020-04-25 | ···················· | ···················· | ···················· | ···················· | ···················· | ···················· | |
| E | 918 hCoV-19/Oman/C-15758/2020\|EPI_ISL_491141\|2020-05-0 | ···················· | ···················· | ···················· | ···················· | ···················· | ···················· | |
| E | 919 hCoV-19/Oman/C-6187/2020\|EPI_ISL_491142\|2020-04-25 | ···················· | ···················· | ···················· | ···················· | ···················· | ···················· | |
| E | 920 hCoV-19/Oman/C-3240/2020\|EPI_ISL_491143\|2020-04-22 | ···················· | ···················· | ···················· | ···················· | ···················· | ···················· | |
| E | 921 hCoV-19/Oman/20762/2020\|EPI_ISL_491144\|2020-04-13 | ···················· | ···················· | ···················· | ···················· | ···················· | ···················· | |
| E | 922 hCoV-19/Oman/B-7106/2020\|EPI_ISL_491145\|2020-05-06 | ···················· | ···················· | ···················· | ···················· | ······A············· | ···················· | |
| E | 923 hCoV-19/Oman/C-6069/2020\|EPI_ISL_491146\|2020-04-26 | ···················· | ···················· | ···················· | ···················· | ···················· | ···················· | |
| E | 924 hCoV-19/Oman/12096/2020\|EPI_ISL_491147\|2020-04-08 | ···················· | ···················· | ···················· | ···················· | ···················· | ···················· | |
| E | 925 hCoV-19/Oman/20751/2020\|EPI_ISL_491148\|2020-04-13 | ···················· | ···················· | ···················· | ···················· | ···················· | ···················· | |
| E | 926 hCoV-19/Oman/14233/2020\|EPI_ISL_491149\|2020-04-10 | ···················· | ·A·················· | ···················· | ···················· | ···················· | ···A················ | |
| E | 927 hCoV-19/Oman/14251/2020\|EPI_ISL_491150\|2020-04-10 | ···················· | ·A·················· | ···················· | ···················· | ···················· | ···A················ | |
| E | 928 hCoV-19/Oman/24774/2020\|EPI_ISL_491151\|2020-04-17 | ···················· | ·A·················· | ···················· | ···················· | ···················· | ···A················ | |
| E | 929 hCoV-19/Oman/16908/2020\|EPI_ISL_491152\|2020-04-12 | ···················· | ·A·················· | ···················· | ···················· | ···················· | ···A················ | |
| E | 930 hCoV-19/Oman/C-16516/2020\|EPI_ISL_491153\|2020-05-0 | ···················· | ···················· | ···················· | ···················· | ···················· | ···················· | |
| E | 931 hCoV-19/Oman/6990/2020\|EPI_ISL_491154\|2020-03-29 | ···················· | ···················· | ···················· | ···················· | ···················· | ···················· | |
| E | 932 hCoV-19/Oman/11374/2020\|EPI_ISL_491155\|2020-04-07 | ···················· | ·A·················· | ···················· | ···················· | ···················· | ···A················ | |
| E | 933 hCoV-19/Oman/14235/2020\|EPI_ISL_491156\|2020-04-10 | ···················· | ·A·················· | ···················· | ···················· | ···················· | ···A················ | |
| E | 934 hCoV-19/Oman/14264/2020\|EPI_ISL_491157\|2020-04-10 | ···················· | ·A·················· | ···················· | ···················· | ···················· | ···A················ | |
| E | 935 hCoV-19/Oman/20616/2020\|EPI_ISL_491158\|2020-04-15 | ···················· | ·A·················· | ···················· | ···················· | ···················· | ···A················ | |
| E | 936 hCoV-19/Oman/15253/2020\|EPI_ISL_491159\|2020-04-12 | ···················· | ·A·················· | ···················· | ···················· | ···················· | ···A················ | |
| E | 937 hCoV-19/Oman/14237/2020\|EPI_ISL_491160\|2020-04-10 | ···················· | ·A·················· | ···················· | ···················· | ···················· | ···A················ | |
| E | 938 hCoV-19/Oman/11467/2020\|EPI_ISL_491161\|2020-04-06 | ···················· | ···················· | ···················· | ···················· | ···················· | ···················· | |
| E | 939 hCoV-19/Oman/C-2673/2020\|EPI_ISL_491162\|2020-04-22 | ···················· | ···················· | ···················· | ···················· | ···················· | ···················· | |
| E | 940 hCoV-19/Oman/C-2826/2020\|EPI_ISL_491163\|2020-04-22 | ···················· | ···················· | ···················· | ···················· | ···················· | ···················· | |
| E | 941 hCoV-19/Oman/C-5401/2020\|EPI_ISL_491164\|2020-04-24 | ···················· | ···················· | ···················· | ···················· | ···················· | ···················· | |
| E | 942 hCoV-19/Oman/C-5806/2020\|EPI_ISL_491165\|2020-04-24 | ···················· | ···················· | ···················· | ···················· | ···················· | ···················· | |
| E | 943 hCoV-19/Oman/B-3125/2020\|EPI_ISL_491166\|2020-04-26 | ···················· | ···················· | ···················· | ···················· | ···················· | ···················· | |
| E | 944 hCoV-19/Oman/20099/2020\|EPI_ISL_491167\|2020-04-15 | ···················· | ···················· | ···················· | ···················· | ···················· | ···················· | |
| E | 945 hCoV-19/Oman/C-1186/2020\|EPI_ISL_491168\|2020-04-21 | ···················· | ···················· | ···················· | ···················· | ···················· | ···················· | |
| E | 946 hCoV-19/Oman/B-6050/2020\|EPI_ISL_491169\|2020-05-03 | ···················· | ···················· | ···················· | ···················· | ···················· | ···················· | |
| E | 947 hCoV-19/Oman/C-11911/2020\|EPI_ISL_491170\|2020-05-0 | ···················· | ···················· | ···················· | ···················· | ···················· | ···················· | |
| E | 948 hCoV-19/Oman/C-3067/2020\|EPI_ISL_491171\|2020-04-22 | ···················· | ···················· | ···················· | ···················· | ···················· | ···················· | |
| E | 949 hCoV-19/Peru/LIM-INS-019/2020\|EPI_ISL_491172\|2020- | ···················· | ···················· | ···················· | ···················· | ···················· | ···················· | |
| E | 950 hCoV-19/Portugal/IGC2885/2020\|EPI_ISL_491175\|2020- | ···················· | ···················· | ···················· | ···················· | ···················· | ···················· | |
| E | 951 hCoV-19/Portugal/IGC3008/2020\|EPI_ISL_491183\|2020- | ···················· | ···················· | ···················· | ···················· | ···················· | ···················· | |
| E | 952 hCoV-19/Portugal/IGC3009/2020\|EPI_ISL_491184\|2020- | ···················· | ···················· | ···················· | ···················· | ···················· | NNN················· | 1 |
| E | 953 hCoV-19/Portugal/IGC3012/2020\|EPI_ISL_491187\|2020- | ···················· | ···················· | ···················· | ···················· | ···················· | ···················· | |
| E | 954 hCoV-19/Portugal/IGC3484/2020\|EPI_ISL_491189\|2020- | ···················· | ···················· | ···················· | ···················· | ···················· | ···················· | |
| E | 955 hCoV-19/England/BRIS-18539BD/2020\|EPI_ISL_492430\|2 | ···················· | ····A··············· | ···················· | ···················· | ···················· | ···················· | |
| E | 956 hCoV-19/England/BRIS-1853FCA/2020\|EPI_ISL_492433\|2 | ···················· | ···················· | ···················· | ···················· | ···················· | ···················· | |
| E | 957 hCoV-19/England/BRIS-185447F/2020\|EPI_ISL_492434\|2 | ···················· | ···················· | ···················· | ···················· | ···················· | ···················· | |
| E | 958 hCoV-19/England/BRIS-1854424/2020\|EPI_ISL_492437\|2 | ···················· | ···················· | ···················· | ···················· | ···················· | ···················· | |
| E | 959 hCoV-19/England/BRIS-18537C2/2020\|EPI_ISL_492439\|2 | ···················· | ···················· | ···················· | ···················· | ···················· | ···················· | |
| E | 960 hCoV-19/England/BRIS-18542FD/2020\|EPI_ISL_492440\|2 | ···················· | ···················· | ···················· | ···················· | ···················· | ···················· | |
| E | 961 hCoV-19/England/BRIS-18539EA/2020\|EPI_ISL_492441\|2 | ···················· | ···················· | ···················· | ···················· | ···················· | ···················· | |
| E | 962 hCoV-19/England/BRIS-1853DC0/2020\|EPI_ISL_492442\|2 | ···················· | ···················· | ···················· | ···················· | ···················· | ···················· | |
| E | 963 hCoV-19/England/BRIS-18531D3/2020\|EPI_ISL_492444\|2 | ···················· | ···················· | ···················· | ···················· | ···················· | ···················· | |
| E | 964 hCoV-19/England/BRIS-185422A/2020\|EPI_ISL_492445\|2 | ···················· | ···················· | ···················· | ···················· | ···················· | ···················· | |
| E | 965 hCoV-19/England/BRIS-18543FA/2020\|EPI_ISL_492447\|2 | ···················· | ···················· | ···················· | ···················· | ···················· | ···················· | |
| E | 966 hCoV-19/England/BRIS-1853FF7/2020\|EPI_ISL_492448\|2 | ···················· | ···················· | ···················· | ···················· | ···················· | ···················· | |
| E | 967 hCoV-19/England/BRIS-18542DF/2020\|EPI_ISL_492449\|2 | ···················· | ···················· | ···················· | ···················· | ···················· | ···················· | |
| E | 968 hCoV-19/England/NORT-29950B/2020\|EPI_ISL_492450\|20 | ···················· | ···················· | ···················· | ···················· | ···················· | ···················· | |
| E | 969 hCoV-19/England/NORT-2994A4/2020\|EPI_ISL_492451\|20 | ···················· | ···················· | ···················· | ···················· | ···················· | ···················· | |
| E | 970 hCoV-19/England/NORT-29AAFC/2020\|EPI_ISL_492452\|20 | ···················· | ···················· | ···················· | ···················· | ···················· | ···················· | |
| E | 971 hCoV-19/England/NORT-29A8E3/2020\|EPI_ISL_492453\|20 | ···················· | ···················· | ···················· | ···················· | ···················· | ···················· | |

| | | | | | | | |
|---|---|---|---|---|---|---|---|
| E | 972 hCoV-19/England/NORT-29AB71/2020\|EPI_ISL_492454\|20 | ···················· | ···················· | ····················· | ···················· | ···················· | ···················· |
| E | 973 hCoV-19/England/NORT-299477/2020\|EPI_ISL_492455\|20 | ···················· | ···················· | ····················· | ···················· | ···················· | ···················· |
| E | 974 hCoV-19/England/NORT-29A8B6/2020\|EPI_ISL_492456\|20 | ···················· | ···················· | ····················· | ···················· | ···················· | ···················· |
| E | 975 hCoV-19/England/NORT-29ABF9/2020\|EPI_ISL_492458\|20 | ···················· | ···················· | ····················· | ···················· | ···················· | ···················· |
| E | 976 hCoV-19/England/NORT-2995A1/2020\|EPI_ISL_492459\|20 | ···················· | ···················· | ····················· | ···················· | ···················· | ···················· |
| E | 977 hCoV-19/England/NORT-29AD11/2020\|EPI_ISL_492461\|20 | ···················· | ···················· | ····················· | ···················· | ···················· | ···················· |
| E | 978 hCoV-19/England/NORT-29AE2D/2020\|EPI_ISL_492462\|20 | ···················· | ···················· | ····················· | ···················· | ···················· | ···················· |
| E | 979 hCoV-19/England/NORT-29986C/2020\|EPI_ISL_492463\|20 | ···················· | ···················· | ····················· | ···················· | ···················· | ···················· |
| E | 980 hCoV-19/England/NORT-29A8A7/2020\|EPI_ISL_492464\|20 | ···················· | ···················· | ····················· | ···················· | ···················· | ···················· |
| E | 981 hCoV-19/England/NORT-299B45/2020\|EPI_ISL_492465\|20 | ···················· | ···················· | ····················· | ···················· | ···················· | ···················· |
| E | 982 hCoV-19/England/NORT-299AB1/2020\|EPI_ISL_492466\|20 | ···················· | ···················· | ····················· | ···················· | ···················· | ···················· |
| E | 983 hCoV-19/England/NORT-29AD4E/2020\|EPI_ISL_492469\|20 | ···················· | ···················· | ····················· | ···················· | ···················· | ···················· |
| E | 984 hCoV-19/England/NORT-299459/2020\|EPI_ISL_492470\|20 | ···················· | ···················· | ····················· | ···················· | ···················· | ···················· |
| E | 985 hCoV-19/England/NORT-2994D1/2020\|EPI_ISL_492471\|20 | ···················· | ···················· | ····················· | ···················· | ···················· | ···················· |
| E | 986 hCoV-19/England/NORT-29AB80/2020\|EPI_ISL_492472\|20 | ···················· | ···················· | ····················· | ···················· | ···················· | ···················· |
| E | 987 hCoV-19/England/NORT-299635/2020\|EPI_ISL_492473\|20 | ···················· | ···················· | ····················· | ···················· | ···················· | ···················· |
| E | 988 hCoV-19/England/NORT-29A8D4/2020\|EPI_ISL_492474\|20 | ···················· | ···················· | ····················· | ···················· | ···················· | ···················· |
| E | 989 hCoV-19/England/NORT-299D30/2020\|EPI_ISL_492475\|20 | ···················· | ···················· | ····················· | ···················· | ···················· | ···················· |
| E | 990 hCoV-19/England/NORT-2995CF/2020\|EPI_ISL_492476\|20 | ···················· | ···················· | ····················· | ···················· | ···················· | ···················· |
| E | 991 hCoV-19/England/NORT-299A93/2020\|EPI_ISL_492477\|20 | ···················· | ···················· | ····················· | ···················· | ···················· | ···················· |
| E | 992 hCoV-19/England/NORT-299AA2/2020\|EPI_ISL_492478\|20 | ···················· | ···················· | ····················· | ···················· | ···················· | ···················· |
| E | 993 hCoV-19/England/NORT-2994FF/2020\|EPI_ISL_492479\|20 | ···················· | ···················· | ····················· | ···················· | ···················· | ···················· |
| E | 994 hCoV-19/England/NORT-299529/2020\|EPI_ISL_492480\|20 | ···················· | ···················· | ····················· | ···················· | ···················· | ···················· |
| E | 995 hCoV-19/England/NORT-299486/2020\|EPI_ISL_492481\|20 | ···················· | ···················· | ····················· | ···················· | ···················· | ···················· |
| E | 996 hCoV-19/England/NORT-299574/2020\|EPI_ISL_492482\|20 | ···················· | ···················· | ····················· | ···················· | ···················· | ···················· |
| E | 997 hCoV-19/England/NORT-29ABEA/2020\|EPI_ISL_492483\|20 | ···················· | ···················· | ····················· | ···················· | ···················· | ···················· |
| E | 998 hCoV-19/England/NORT-29AA0B/2020\|EPI_ISL_492484\|20 | ···················· | ···················· | ····················· | ···················· | ···················· | ···················· |
| E | 999 hCoV-19/England/NORT-2994C2/2020\|EPI_ISL_492485\|20 | ···················· | ···················· | ····················· | ···················· | ···················· | ···················· |
| E | 1000 hCoV-19/England/NORT-299592/2020\|EPI_ISL_492487\|20 | ···················· | ···················· | ····················· | ···················· | ···················· | ···················· |
| E | 1001 hCoV-19/England/NORT-299565/2020\|EPI_ISL_492488\|20 | ···················· | ···················· | ····················· | ···················· | ···················· | ···················· |
| E | 1002 hCoV-19/England/NORT-299495/2020\|EPI_ISL_492489\|20 | ···················· | ···················· | ····················· | ···················· | ···················· | ···················· |
| E | 1003 hCoV-19/England/NORT-29A9FF/2020\|EPI_ISL_492490\|20 | ···················· | ···················· | ····················· | ···················· | ···················· | ···················· |
| E | 1004 hCoV-19/England/NORT-29944A/2020\|EPI_ISL_492491\|20 | ···················· | ···················· | ····················· | ···················· | ···················· | ···················· |
| E | 1005 hCoV-19/England/NORT-29AC7E/2020\|EPI_ISL_492492\|20 | ···················· | ···················· | ····················· | ···················· | ···················· | ···················· |
| E | 1006 hCoV-19/England/NORT-29ADC6/2020\|EPI_ISL_492493\|20 | ···················· | ···················· | ····················· | ···················· | ···················· | ···················· |
| E | 1007 hCoV-19/England/LOND-D7EAE/2020\|EPI_ISL_492496\|202 | ···················· | ···················· | ····················· | ···················· | ···················· | ···················· |
| E | 1008 hCoV-19/England/LOND-D7BC5/2020\|EPI_ISL_492498\|202 | ···················· | ···················· | ····················· | ···················· | K··················· | ···················· |
| E | 1025 hCoV-19/USA/WI-UW-428/2020\|EPI_ISL_480410\|2020-05- | ···················· | ···················· | ····················· | ···················· | ··············N····· | ·NN················· |
| E | 1026 hCoV-19/USA/WI-UW-429/2020\|EPI_ISL_480411\|2020-05- | ···················· | ···················· | ····················· | ···················· | ···················· | ···················· |
| E | 1027 hCoV-19/USA/WI-UW-430/2020\|EPI_ISL_480412\|2020-05- | ···················· | ···················· | ····················· | ···················· | ···················· | ···················· |
| E | 1028 hCoV-19/USA/WI-UW-431/2020\|EPI_ISL_480413\|2020-05- | ···················· | ···················· | ····················· | ···················· | ···················· | ···················· |
| E | 1029 hCoV-19/Bangladesh/BCSIR-NILMRC-130/2020\|EPI_ISL_4 | ···················· | ···················· | ····················· | ···················· | ···················· | ···················· |
| E | 1030 hCoV-19/Bangladesh/BCSIR-NILMRC-131/2020\|EPI_ISL_4 | ···················· | ···················· | ····················· | ···················· | ···················· | ···················· |
| E | 1031 hCoV-19/Bangladesh/BCSIR-NILMRC-132/2020\|EPI_ISL_4 | ···················· | ···················· | ····················· | ···················· | ···················· | ···················· |
| E | 1032 hCoV-19/Bangladesh/BCSIR-NILMRC-141/2020\|EPI_ISL_4 | ···················· | ···················· | ····················· | ···················· | ···················· | ···················· |
| E | 1033 hCoV-19/Bangladesh/BCSIR-NILMRC-144/2020\|EPI_ISL_4 | ···················· | ···················· | ····················· | ···················· | ···················· | ···················· |
| E | 1034 hCoV-19/Bangladesh/BCSIR-NILMRC-133/2020\|EPI_ISL_4 | ···················· | ···················· | ·········T··········· | ···················· | ···················· | ···················· |
| E | 1035 hCoV-19/Bangladesh/BCSIR-NILMRC-135/2020\|EPI_ISL_4 | ···················· | ···················· | ····················· | ···················· | ···················· | ···················· |
| E | 1036 hCoV-19/Bangladesh/BCSIR-NILMRC-137/2020\|EPI_ISL_4 | ···················· | ···················· | ····················· | ···················· | ···················· | ···················· |
| E | 1037 hCoV-19/Bangladesh/BCSIR-NILMRC-138/2020\|EPI_ISL_4 | ···················· | ···················· | ····················· | ···················· | ···················· | ···················· |
| E | 1038 hCoV-19/Bangladesh/BCSIR-NILMRC-139/2020\|EPI_ISL_4 | ···················· | ···················· | ····················· | ···················· | ···················· | ···················· |
| E | 1039 hCoV-19/Bangladesh/BCSIR-NILMRC-145/2020\|EPI_ISL_4 | ···················· | ···················· | ····················· | ···················· | ···················· | ···················· |
| E | 1040 hCoV-19/Bangladesh/BCSIR-NILMRC-146/2020\|EPI_ISL_4 | ···················· | ···················· | ····················· | ···················· | ···················· | ···················· |
| E | 1041 hCoV-19/Bangladesh/BCSIR-NILMRC-150/2020\|EPI_ISL_4 | ···················· | ···················· | ····················· | ···················· | ···················· | ···················· |
| E | 1042 hCoV-19/Bangladesh/BCSIR-NILMRC-151/2020\|EPI_ISL_4 | ···················· | ···················· | ····················· | ···················· | ···················· | ···················· |
| E | 1043 hCoV-19/Bangladesh/BCSIR-NILMRC-152/2020\|EPI_ISL_4 | ···················· | ···················· | ····················· | ···················· | ···················· | ···················· |
| E | 1044 hCoV-19/Bangladesh/BCSIR-NILMRC-153/2020\|EPI_ISL_4 | ···················· | ···················· | ····················· | ···················· | ···················· | ···················· |
| E | 1045 hCoV-19/Bangladesh/BCSIR-NILMRC-125/2020\|EPI_ISL_4 | ···················· | ···················· | ····················· | ···················· | ···················· | ···················· |
| E | 1046 hCoV-19/Bangladesh/BCSIR-NILMRC-126/2020\|EPI_ISL_4 | ···················· | ···················· | ····················· | ···················· | ···················· | ···················· |
| E | 1047 hCoV-19/Bangladesh/BCSIR-NILMRC-154/2020\|EPI_ISL_4 | ···················· | ···················· | ····················· | ···················· | ···················· | ···················· |
| E | 1048 hCoV-19/Bangladesh/BCSIR-NILMRC-156/2020\|EPI_ISL_4 | ···················· | ···················· | ····················· | ···················· | ···················· | ···················· |
| E | 1049 hCoV-19/Bangladesh/BCSIR-NILMRC-158/2020\|EPI_ISL_4 | ···················· | ·A·················· | ····················· | ···················· | ···················· | ··A················· |
| E | 1050 hCoV-19/Bangladesh/BCSIR-NILMRC-168/2020\|EPI_ISL_4 | ···················· | ···················· | ····················· | ···················· | ···················· | ···················· |
| E | 1051 hCoV-19/Bangladesh/BCSIR-NILMRC-172/2020\|EPI_ISL_4 | ···················· | ···················· | ····················· | ···················· | ···················· | ···················· |
| E | 1052 hCoV-19/Finland/9A85S12/2020\|EPI_ISL_481740\|2020-0 | ···················· | ···················· | ····················· | ···················· | ···················· | ···················· |
| E | 1053 hCoV-19/Canada/NB_243/2020\|EPI_ISL_481742\|2020-03- | ···················· | ···················· | ····················· | ···················· | ···················· | ···················· |
| E | 1054 hCoV-19/Canada/NB_245/2020\|EPI_ISL_481743\|2020-03- | ···················· | ···················· | ····················· | ···················· | ···················· | ···················· |
| E | 1055 hCoV-19/Canada/NB_246/2020\|EPI_ISL_481744\|2020-03- | ···················· | ···················· | ····················· | ···················· | ···················· | ···················· |
| E | 1056 hCoV-19/Canada/NB_247/2020\|EPI_ISL_481745\|2020-03- | ···················· | ···················· | ····················· | ···················· | ···················· | ···················· |
| E | 1057 hCoV-19/Canada/NB_248/2020\|EPI_ISL_481746\|2020-04- | ···················· | ···················· | ····················· | ···················· | ···················· | ···················· |
| E | 1058 hCoV-19/Canada/NB_249/2020\|EPI_ISL_481747\|2020-03- | ···················· | ···················· | ····················· | ···················· | ···················· | ···················· |
| E | 1059 hCoV-19/Canada/NB_250/2020\|EPI_ISL_481748\|2020-04- | ···················· | ···················· | ····················· | ···················· | ···················· | ···················· |
| E | 1060 hCoV-19/Canada/NB_252/2020\|EPI_ISL_481749\|2020-04- | ···················· | ···················· | ····················· | ···················· | ···················· | ···················· |
| E | 1061 hCoV-19/Canada/NB_257/2020\|EPI_ISL_481750\|2020-04- | ···················· | ···················· | ····················· | ···················· | ···················· | ···················· |
| E | 1062 hCoV-19/Canada/NB_260/2020\|EPI_ISL_481751\|2020-04- | ···················· | ···················· | ····················· | ···················· | ···················· | ···················· |
| E | 1063 hCoV-19/Canada/NB_261/2020\|EPI_ISL_481752\|2020-04- | ···················· | ···················· | ····················· | ···················· | ···················· | ···················· |
| E | 1064 hCoV-19/Canada/NB_263/2020\|EPI_ISL_481753\|2020-04- | ···················· | ···················· | ····················· | ···················· | ···················· | ···················· |
| E | 1065 hCoV-19/Canada/NB_264/2020\|EPI_ISL_481754\|2020-04- | ···················· | ···················· | ····················· | ···················· | ···················· | ···················· |
| E | 1066 hCoV-19/Canada/NB_270/2020\|EPI_ISL_481755\|2020-04- | ···················· | ···················· | ····················· | ···················· | ···················· | ···················· |
| E | 1067 hCoV-19/Canada/NB_271/2020\|EPI_ISL_481756\|2020-04- | ···················· | ···················· | ····················· | ···················· | ···················· | ···················· |

| | | | | | | | | |
|---|---|---|---|---|---|---|---|---|
| E | 1068 | hCoV-19/Canada/NB_274/2020\|EPI_ISL_481757\|2020-04- | .................... | .................... | .................... | .................... | .................... | .................... |
| E | 1069 | hCoV-19/Canada/NB_277/2020\|EPI_ISL_481758\|2020-04- | .................... | .................... | .................... | .................... | .................... | .................... |
| E | 1070 | hCoV-19/England/BRIS-12DB1A/2020\|EPI_ISL_481768\|20 | .................... | .................... | .................... | .................... | .................... | .................... |
| E | 1071 | hCoV-19/England/BRIS-12D6EC/2020\|EPI_ISL_481769\|20 | .................... | .................... | .................... | .................... | .................... | .................... |
| E | 1072 | hCoV-19/England/BRIS-12D215/2020\|EPI_ISL_481770\|20 | .................... | .................... | .................... | .................... | .................... | .................... |
| E | 1073 | hCoV-19/England/BRIS-12CC72/2020\|EPI_ISL_481771\|20 | .................... | .................... | .................... | .................... | .................... | .................... |
| E | 1074 | hCoV-19/England/BRIS-12CEB8/2020\|EPI_ISL_481772\|20 | .................... | .................... | .................... | .................... | .................... | .................... |
| E | 1075 | hCoV-19/England/BRIS-12DA0E/2020\|EPI_ISL_481773\|20 | .................... | .................... | .................... | .................... | .................... | .................... |
| E | 1076 | hCoV-19/England/BRIS-12CD06/2020\|EPI_ISL_481774\|20 | .................... | .................... | .................... | .................... | .................... | .................... |
| E | 1077 | hCoV-19/England/BRIS-12D86E/2020\|EPI_ISL_481775\|20 | .................... | .................... | .................... | .................... | .................... | .................... |
| E | 1078 | hCoV-19/England/BRIS-12D94D/2020\|EPI_ISL_481776\|20 | .................... | .................... | .................... | .................... | .................... | .................... |
| E | 1079 | hCoV-19/England/BRIS-12D989/2020\|EPI_ISL_481777\|20 | .................... | .................... | .................... | .................... | .................... | .................... |
| E | 1080 | hCoV-19/England/BRIS-12D163/2020\|EPI_ISL_481779\|20 | .................... | .................... | .................... | .................... | .................... | .................... |
| E | 1081 | hCoV-19/USA/IL-UW-625/2020\|EPI_ISL_485000\|2020-06- | .................... | .................... | .................... | .................... | .................... | .................... |
| E | 1082 | hCoV-19/South Korea/A2/2020\|EPI_ISL_485001\|2020-03 | .................... | .................... | .................... | .................... | .................... | .................... |
| E | 1083 | hCoV-19/South Korea/A3/2020\|EPI_ISL_485002\|2020-03 | .................... | .................... | .................... | .................... | .................... | .................... |
| E | 1084 | hCoV-19/USA/LA-GBCL-26159761/2020\|EPI_ISL_485011\|2 | .................... | .................... | .................... | .................... | .................... | .................... |
| E | 1085 | hCoV-19/USA/LA-GBCL-26159762/2020\|EPI_ISL_485012\|2 | .................... | .................... | .................... | .................... | .................... | .................... |
| E | 1086 | hCoV-19/USA/LA-GBCL-26492443/2020\|EPI_ISL_485026\|2 | .................... | .................... | .................... | .................... | .................... | .................... |
| E | 1087 | hCoV-19/USA/LA-GBCL-26492444/2020\|EPI_ISL_485027\|2 | .................... | .................... | .................... | .................... | .................... | .................... |
| E | 1088 | hCoV-19/USA/LA-GBCL-26492449/2020\|EPI_ISL_485029\|2 | .................... | .................... | .................... | .................... | .................... | .................... |
| E | 1089 | hCoV-19/USA/LA-GBCL-26492453/2020\|EPI_ISL_485031\|2 | .................... | .................... | .................... | .................... | .................... | .................... |
| E | 1090 | hCoV-19/USA/LA-GBCL-26492454/2020\|EPI_ISL_485032\|2 | .................... | .................... | .................... | .................... | .................... | .................... |
| E | 1091 | hCoV-19/USA/LA-GBCL-26492456/2020\|EPI_ISL_485034\|2 | .................... | .................... | .................... | .................... | .................... | .................... |
| E | 1092 | hCoV-19/USA/LA-GBCL-26492457/2020\|EPI_ISL_485035\|2 | .................... | .................... | .................... | .................... | .................... | .................... |
| E | 1093 | hCoV-19/USA/LA-GBCL-26492461/2020\|EPI_ISL_485038\|2 | .................... | .................... | .................... | .................... | .................... | .................... |
| E | 1094 | hCoV-19/USA/LA-GBCL-26492465/2020\|EPI_ISL_485040\|2 | .................... | .................... | .................... | .................... | .................... | .................... |
| E | 1095 | hCoV-19/USA/LA-GBCL-26492469/2020\|EPI_ISL_485041\|2 | .................... | .................... | .................... | .................... | .................... | .................... |
| E | 1096 | hCoV-19/USA/LA-GBCL-26492470/2020\|EPI_ISL_485042\|2 | .................... | .................... | .................... | .................... | .................... | .................... |
| E | 1097 | hCoV-19/USA/LA-GBCL-26492473/2020\|EPI_ISL_485043\|2 | .................... | .................... | .................... | .................... | .................... | .................... |
| E | 1098 | hCoV-19/USA/LA-GBCL-26492479/2020\|EPI_ISL_485045\|2 | .................... | .................... | .................... | .................... | .................... | .................... |
| E | 1099 | hCoV-19/USA/LA-GBCL-26492480/2020\|EPI_ISL_485046\|2 | .................... | .................... | .................... | .................... | .................... | .................... |
| E | 1100 | hCoV-19/USA/LA-GBCL-26492496/2020\|EPI_ISL_485052\|2 | .................... | .................... | .................... | .................... | .................... | .................... |
| E | 1101 | hCoV-19/USA/LA-GBCL-26492498/2020\|EPI_ISL_485054\|2 | .................... | .................... | .................... | .................... | .................... | .................... |
| E | 1102 | hCoV-19/USA/LA-GBCL-26492499/2020\|EPI_ISL_485055\|2 | .................... | .................... | .................... | .................... | .................... | .................... |
| E | 1103 | hCoV-19/USA/LA-GBCL-26492500/2020\|EPI_ISL_485056\|2 | .................... | .................... | .................... | .................... | .................... | .................... |
| E | 1104 | hCoV-19/USA/LA-GBCL-26492502/2020\|EPI_ISL_485057\|2 | .................... | .................... | .................... | .................... | .................... | .................... |
| E | 1105 | hCoV-19/USA/LA-GBCL-26492505/2020\|EPI_ISL_485059\|2 | .................... | .................... | .................... | .................... | .................... | .................... |
| E | 1106 | hCoV-19/USA/LA-GBCL-26492506/2020\|EPI_ISL_485060\|2 | .................... | .................... | .................... | .................... | .................... | .................... |
| E | 1107 | hCoV-19/USA/LA-GBCL-26492512/2020\|EPI_ISL_485062\|2 | .................... | .................... | .................... | .................... | .................... | .................... |
| E | 1108 | hCoV-19/USA/LA-GBCL-26492514/2020\|EPI_ISL_485063\|2 | .................... | .................... | .................... | .................... | .................... | .................... |
| E | 1109 | hCoV-19/USA/LA-GBCL-26512989/2020\|EPI_ISL_485065\|2 | .................... | .................... | .................... | .................... | .................... | .................... |
| E | 1110 | hCoV-19/USA/LA-GBCL-26512997/2020\|EPI_ISL_485067\|2 | .................... | .................... | .................... | .................... | .................... | .................... |
| E | 1111 | hCoV-19/USA/LA-GBCL-26512999/2020\|EPI_ISL_485068\|2 | .................... | .................... | .................... | .................... | .................... | .................... |
| E | 1112 | hCoV-19/USA/LA-GBCL-26513004/2020\|EPI_ISL_485069\|2 | .................... | .................... | .................... | .................... | .................... | .................... |
| E | 1113 | hCoV-19/USA/LA-GBCL-26513006/2020\|EPI_ISL_485070\|2 | .................... | .................... | .................... | .................... | .................... | .................... |
| E | 1114 | hCoV-19/USA/LA-GBCL-26513008/2020\|EPI_ISL_485071\|2 | .................... | .................... | .................... | .................... | .................... | .................... |
| E | 1115 | hCoV-19/USA/LA-GBCL-26513012/2020\|EPI_ISL_485072\|2 | .................... | .................... | .................... | .................... | .................... | .................... |
| E | 1116 | hCoV-19/USA/LA-GBCL-26513013/2020\|EPI_ISL_485073\|2 | .................... | .................... | .................... | .................... | .................... | .................... |
| E | 1117 | hCoV-19/USA/LA-GBCL-26513022/2020\|EPI_ISL_485076\|2 | .................... | .................... | .................... | .................... | .................... | .................... |
| E | 1118 | hCoV-19/USA/LA-GBCL-26513023/2020\|EPI_ISL_485077\|2 | .................... | .................... | .................... | .................... | .................... | .................... |
| E | 1119 | hCoV-19/USA/LA-GBCL-26513025/2020\|EPI_ISL_485078\|2 | .................... | .................... | .................... | .................... | .................... | .................... |
| E | 1120 | hCoV-19/USA/LA-GBCL-26513028/2020\|EPI_ISL_485079\|2 | .................... | .................... | .................... | .................... | .................... | .................... |
| E | 1121 | hCoV-19/USA/LA-GBCL-26513029/2020\|EPI_ISL_485080\|2 | .................... | .................... | .................... | .................... | .................... | .................... |
| E | 1122 | hCoV-19/USA/LA-GBCL-26513032/2020\|EPI_ISL_485081\|2 | .................... | .................... | .................... | .................... | .................... | .................... |
| E | 1123 | hCoV-19/USA/LA-GBCL-26513034/2020\|EPI_ISL_485082\|2 | .................... | .................... | .................... | .................... | .................... | .................... |
| E | 1124 | hCoV-19/USA/LA-GBCL-26513035/2020\|EPI_ISL_485083\|2 | .................... | .................... | .................... | .................... | .................... | .................... |
| E | 1125 | hCoV-19/USA/LA-GBCL-26513036/2020\|EPI_ISL_485084\|2 | .................... | .................... | .................... | .................... | .................... | .................... |
| E | 1126 | hCoV-19/USA/LA-GBCL-26513042/2020\|EPI_ISL_485086\|2 | .................... | .................... | .................... | .................... | .................... | .................... |
| E | 1127 | hCoV-19/USA/LA-GBCL-26513049/2020\|EPI_ISL_485089\|2 | .................... | .................... | .................... | .................... | .................... | .................... |
| E | 1128 | hCoV-19/USA/LA-GBCL-26513050/2020\|EPI_ISL_485090\|2 | .................... | .................... | .................... | .................... | .................... | .................... |
| E | 1129 | hCoV-19/USA/LA-GBCL-26513055/2020\|EPI_ISL_485091\|2 | .................... | .................... | .................... | .................... | .................... | .................... |
| E | 1130 | hCoV-19/USA/LA-GBCL-26513059/2020\|EPI_ISL_485094\|2 | .................... | .................... | .................... | .................... | .................... | .................... |
| E | 1131 | hCoV-19/USA/LA-GBCL-26513062/2020\|EPI_ISL_485095\|2 | .................... | .................... | .................... | .................... | .................... | .................... |
| E | 1132 | hCoV-19/USA/LA-GBCL-26513063/2020\|EPI_ISL_485096\|2 | .................... | .................... | .................... | .................... | .................... | .................... |
| E | 1133 | hCoV-19/USA/LA-GBCL-26513064/2020\|EPI_ISL_485097\|2 | .................... | .................... | .................... | .................... | .................... | .................... |
| E | 1134 | hCoV-19/USA/LA-GBCL-26513066/2020\|EPI_ISL_485098\|2 | .................... | .................... | .................... | .................... | .................... | .................... |
| E | 1135 | hCoV-19/USA/CA-CZB-2069/2020\|EPI_ISL_486312\|2020-0 | .................... | .................... | .................... | .................... | .................... | .................... |
| E | 1136 | hCoV-19/USA/CA-CZB-2070/2020\|EPI_ISL_486313\|2020-0 | .................... | .................... | .................... | .................... | .................... | .................... |
| E | 1137 | hCoV-19/USA/CA-CZB-2071/2020\|EPI_ISL_486314\|2020-0 | .................... | .................... | .................... | .................... | .................... | .................... |
| E | 1138 | hCoV-19/USA/CA-CZB-2072/2020\|EPI_ISL_486315\|2020-0 | .................... | .................... | .................... | .................... | .................... | .................... |
| E | 1139 | hCoV-19/USA/CA-CZB-2073/2020\|EPI_ISL_486316\|2020-0 | .................... | .................... | .................... | .................... | .................... | .................... |
| E | 1140 | hCoV-19/USA/CA-CZB-2074/2020\|EPI_ISL_486317\|2020-0 | .................... | .................... | .................... | .................... | .................... | .................... |
| E | 1141 | hCoV-19/USA/CA-CZB-2075/2020\|EPI_ISL_486318\|2020-0 | .................... | .................... | .................... | .................... | .................... | .................... |
| E | 1142 | hCoV-19/USA/CA-CZB-2076/2020\|EPI_ISL_486319\|2020-0 | .................... | .................... | .................... | .................... | .................... | .................... |
| E | 1143 | hCoV-19/USA/CA-CZB-2077/2020\|EPI_ISL_486320\|2020-0 | .................... | .................... | .................... | .................... | .................... | .................... |
| E | 1144 | hCoV-19/USA/CA-CZB-2078/2020\|EPI_ISL_486321\|2020-0 | .................... | .................... | .................... | .................... | .................... | .................... |
| E | 1145 | hCoV-19/USA/CA-CZB-2079/2020\|EPI_ISL_486322\|2020-0 | .................... | .................... | .................... | .................... | .................... | .................... |
| E | 1146 | hCoV-19/USA/CA-CZB-2080/2020\|EPI_ISL_486323\|2020-0 | .................... | .................... | .................... | .................... | .................... | .................... |
| E | 1147 | hCoV-19/USA/CA-CZB-2081/2020\|EPI_ISL_486324\|2020-0 | .................... | .................... | .................... | .................... | .................... | .................... |

| | | | | | | | | | |
|---|---|---|---|---|---|---|---|---|---|
| E | 1148 | hCoV-19/USA/CA-CZB-2082/2020\|EPI_ISL_486325\|2020-0 | ···················· | ···················· | ····················· | ···················· | ···················· | ···················· | |
| E | 1149 | hCoV-19/USA/CA-CZB-2083/2020\|EPI_ISL_486326\|2020-0 | ···················· | ···················· | ····················· | ···················· | ···················· | ···················· | |
| E | 1150 | hCoV-19/USA/CA-CZB-2084/2020\|EPI_ISL_486327\|2020-0 | ···················· | ···················· | ····················· | ···················· | ···················· | ···················· | |
| E | 1151 | hCoV-19/USA/CA-CZB-2085/2020\|EPI_ISL_486328\|2020-0 | ···················· | ···················· | ····················· | ···················· | ···················· | ···················· | |
| E | 1152 | hCoV-19/USA/CA-CZB-2086/2020\|EPI_ISL_486329\|2020-0 | ···················· | ···················· | ····················· | ···················· | ···················· | ···················· | |
| E | 1153 | hCoV-19/USA/CA-CZB-2087/2020\|EPI_ISL_486330\|2020-0 | ···················· | ···················· | ····················· | ···················· | ···················· | ···················· | |
| E | 1154 | hCoV-19/USA/CA-CZB-2088/2020\|EPI_ISL_486331\|2020-0 | ···················· | ···················· | ····················· | ···················· | ···················· | ···················· | |
| E | 1155 | hCoV-19/USA/CA-CZB-2089/2020\|EPI_ISL_486332\|2020-0 | ···················· | ···················· | ····················· | ···················· | ···················· | ···················· | |
| E | 1156 | hCoV-19/USA/CA-CZB-2109/2020\|EPI_ISL_486342\|2020-0 | ···················· | ···················· | ····················· | ···················· | ···················· | ···················· | |
| E | 1157 | hCoV-19/USA/CA-CZB-2111/2020\|EPI_ISL_486343\|2020-0 | ···················· | ···················· | ····················· | ···················· | ···················· | ···················· | |
| E | 1158 | hCoV-19/USA/CA-CZB-2113/2020\|EPI_ISL_486344\|2020-0 | ···················· | ···················· | ····················· | ···················· | ···················· | ···················· | |
| E | 1159 | hCoV-19/USA/CA-CZB-2129/2020\|EPI_ISL_486356\|2020-0 | ···················· | ···················· | ····················· | ···················· | ···················· | ···················· | |
| E | 1160 | hCoV-19/USA/CA-CZB-2130/2020\|EPI_ISL_486357\|2020-0 | ···················· | ···················· | ····················· | ···················· | ···················· | ···················· | |
| E | 1161 | hCoV-19/USA/CA-CZB-2131/2020\|EPI_ISL_486358\|2020-0 | ···················· | ···················· | ····················· | ···················· | ···················· | ···················· | |
| E | 1162 | hCoV-19/USA/CA-CZB-2134/2020\|EPI_ISL_486360\|2020-0 | ···················· | ···················· | ····················· | ···················· | ···················· | ···················· | |
| E | 1163 | hCoV-19/USA/CA-CZB-2139/2020\|EPI_ISL_486361\|2020-0 | ···················· | ···················· | ····················· | ···················· | ···················· | ···················· | |
| E | 1164 | hCoV-19/USA/CA-CZB-2140/2020\|EPI_ISL_486362\|2020-0 | ···················· | ···················· | ····················· | ···················· | ···················· | ···················· | |
| E | 1165 | hCoV-19/USA/CA-CZB-2141/2020\|EPI_ISL_486363\|2020-0 | ···················· | ···················· | ····················· | ···················· | ···················· | ···················· | |
| E | 1166 | hCoV-19/USA/CA-CZB-2148/2020\|EPI_ISL_486365\|2020-0 | ···················· | ···················· | ····················· | ···················· | ···················· | ···················· | |
| E | 1167 | hCoV-19/India/KA-nimh-0999/2020\|EPI_ISL_486382\|202 | ···················· | ···················· | ····················· | ···················· | ···················· | ···················· | |
| E | 1168 | hCoV-19/India/KA-nimh-3959/2020\|EPI_ISL_486383\|202 | ···················· | ···················· | ····················· | ···················· | ···················· | ···················· | |
| E | 1169 | hCoV-19/India/KA-nimh-14723/2020\|EPI_ISL_486388\|20 | ···················· | ···················· | ····················· | ···················· | ···················· | ···················· | |
| E | 1170 | hCoV-19/India/KA-nimh-15835/2020\|EPI_ISL_486389\|20 | ···················· | ···················· | ····················· | ···················· | ···················· | ···················· | |
| E | 1171 | hCoV-19/Latvia/025/2020\|EPI_ISL_486390\|2020-05-09 | ···················· | ···················· | ····················· | ···················· | ···················· | ···················· | |
| E | 1172 | hCoV-19/Latvia/026/2020\|EPI_ISL_486391\|2020-05-03 | ···················· | ···················· | ····················· | ···················· | ···················· | ···················· | |
| E | 1173 | hCoV-19/India/KA-nimh-1454/2020\|EPI_ISL_486392\|202 | ···················· | ···················· | ····················· | ···················· | ···················· | ···················· | |
| E | 1174 | hCoV-19/India/KA-nimh-3720/2020\|EPI_ISL_486394\|202 | ···················· | ···················· | ····················· | ···················· | ···················· | ···················· | |
| E | 1175 | hCoV-19/India/KA-nimh-4263/2020\|EPI_ISL_486395\|202 | ···················· | ···················· | ····················· | ···················· | ···················· | ···················· | |
| E | 1176 | hCoV-19/India/KA-nimh-5102/2020\|EPI_ISL_486397\|202 | ···················· | ···················· | ····················· | ···················· | ···················· | ···················· | |
| E | 1177 | hCoV-19/India/KA-nimh-5297/2020\|EPI_ISL_486398\|202 | ···················· | ···················· | ····················· | ···················· | ···················· | ···················· | |
| E | 1178 | hCoV-19/India/KA-nimh-7817/2020\|EPI_ISL_486399\|202 | ···················· | ···················· | ····················· | ···················· | ···················· | ···················· | |
| E | 1179 | hCoV-19/England/LOND-D96E0/2020\|EPI_ISL_487643\|202 | ···················· | ···················· | ····················· | ···················· | ···················· | ···················· | |
| E | 1180 | hCoV-19/England/LOND-D96D1/2020\|EPI_ISL_487646\|202 | ···················· | ···················· | ····················· | ···················· | ···················· | ···················· | |
| E | 1181 | hCoV-19/Scotland/EDB4138/2020\|EPI_ISL_487647\|2020- | ···················· | ···················A | ····················· | ···················· | ···················· | ···················· | 1 |
| E | 1182 | hCoV-19/Scotland/EDB5665/2020\|EPI_ISL_487648\|2020- | ···················· | ···················· | ····················· | ···················· | ···················· | ···················· | |
| E | 1183 | hCoV-19/Scotland/EDB4326/2020\|EPI_ISL_487649\|2020- | ···················· | ···················· | ····················· | ···················· | ···················· | ···················· | |
| E | 1184 | hCoV-19/Scotland/EDB5876/2020\|EPI_ISL_487651\|2020- | ···················· | ···················A | ····················· | ···················· | ···················· | ···················· | 1 |
| E | 1185 | hCoV-19/Scotland/EDB2692/2020\|EPI_ISL_487652\|2020- | ···················· | ···················· | ····················· | ···················· | ···················· | ···················· | |
| E | 1186 | hCoV-19/Scotland/EDB3322/2020\|EPI_ISL_487653\|2020- | ···················· | ···················A | ····················· | ···················· | ···················· | ···················· | 1 |
| E | 1187 | hCoV-19/Scotland/EDB2766/2020\|EPI_ISL_487654\|2020- | ···················· | ···················· | ····················· | ···················· | ···················· | ···················· | |
| E | 1188 | hCoV-19/Scotland/EDB3427/2020\|EPI_ISL_487655\|2020- | ···················· | ···················· | ····················· | ···················· | ···················· | ···················· | |
| E | 1189 | hCoV-19/Scotland/EDB3508/2020\|EPI_ISL_487656\|2020- | ···················· | ···················· | ····················· | ···················· | ···················· | ···················· | |
| E | 1190 | hCoV-19/Scotland/EDB6018/2020\|EPI_ISL_487657\|2020- | ···················· | ···················A | ····················· | ···················· | ···················· | ···················· | 1 |
| E | 1191 | hCoV-19/Scotland/EDB5926/2020\|EPI_ISL_487658\|2020- | ···················· | ···················· | ····················· | ···················· | ···················· | ···················· | |
| E | 1192 | hCoV-19/Scotland/EDB6546/2020\|EPI_ISL_487660\|2020- | ···················· | ···················· | ····················· | ···················· | ···················· | ···················· | |
| E | 1193 | hCoV-19/Scotland/EDB3364/2020\|EPI_ISL_487661\|2020- | ···················· | ···················· | ····················· | ···················· | ···················· | ···················· | |
| E | 1194 | hCoV-19/Scotland/EDB5510/2020\|EPI_ISL_487662\|2020- | ···················· | ···················· | ····················· | ···················· | ···················· | ···················· | |
| E | 1195 | hCoV-19/Scotland/EDB5974/2020\|EPI_ISL_487664\|2020- | ···················· | ···················A | ····················· | ···················· | ···················· | ···················· | 1 |
| E | 1196 | hCoV-19/Scotland/EDB3443/2020\|EPI_ISL_487671\|2020- | ···················· | ···················A | ····················· | ···················· | ···················· | ···················· | 1 |
| E | 1197 | hCoV-19/Scotland/EDB4028/2020\|EPI_ISL_487672\|2020- | ···················· | ···················A | ····················· | ···················· | ···················· | ···················· | 1 |
| E | 1198 | hCoV-19/Scotland/EDB5643/2020\|EPI_ISL_487673\|2020- | ···················· | ·············A······ | ····················· | ···················· | ···················· | ···················· | |
| E | 1199 | hCoV-19/Scotland/EDB5956/2020\|EPI_ISL_487674\|2020- | ···················· | ···················A | ····················· | ···················· | ···················· | ···················· | 1 |
| E | 1200 | hCoV-19/Scotland/EDB3535/2020\|EPI_ISL_487675\|2020- | ···················· | ···················A | ····················· | ···················· | ···················· | ···················· | 1 |
| E | 1201 | hCoV-19/Scotland/EDB3536/2020\|EPI_ISL_487687\|2020- | ···················· | ···················· | ····················· | ···················· | ···················· | ···················· | |
| E | 1202 | hCoV-19/Scotland/EDB3357/2020\|EPI_ISL_487688\|2020- | ···················· | ···················· | ····················· | ···················· | ···················· | ···················· | |
| E | 1203 | hCoV-19/Scotland/EDB2976/2020\|EPI_ISL_487689\|2020- | ···················· | ···················· | ····················· | ···················· | ···················· | ···················· | |
| E | 1204 | hCoV-19/Scotland/EDB5918/2020\|EPI_ISL_487690\|2020- | ···················· | ···················· | ····················· | ···················· | ···················· | ···················· | |
| E | 1205 | hCoV-19/Scotland/EDB5883/2020\|EPI_ISL_487691\|2020- | ···················· | ···················A | ····················· | ···················· | ···················· | ···················· | 1 |
| E | 1206 | hCoV-19/Scotland/EDB3507/2020\|EPI_ISL_487692\|2020- | ···················· | ···················· | ····················· | ···················· | ···················· | ···················· | |
| E | 1207 | hCoV-19/Scotland/EDB4068/2020\|EPI_ISL_487694\|2020- | ···················· | ·A·················· | ····················· | ···················· | ···················· | ···················· | |
| E | 1208 | hCoV-19/Scotland/EDB5638/2020\|EPI_ISL_487695\|2020- | ···················· | ···················· | ····················· | ···················· | ···················· | ···················· | |
| E | 1209 | hCoV-19/Scotland/EDB6312/2020\|EPI_ISL_487696\|2020- | ···················· | ···················A | ····················· | ···················· | ···················· | ···················· | 1 |
| E | 1210 | hCoV-19/Scotland/EDB3435/2020\|EPI_ISL_487697\|2020- | ···················· | ···················· | ····················· | ···················· | ···················· | ···················· | |
| E | 1211 | hCoV-19/Scotland/EDB2774/2020\|EPI_ISL_487698\|2020- | ···················· | ···················· | ····················· | ···················· | ···················· | ···················· | |
| E | 1212 | hCoV-19/Scotland/EDB3097/2020\|EPI_ISL_487699\|2020- | ···················· | ···················· | ····················· | ···················· | ···················· | ···················· | |
| E | 1213 | hCoV-19/Scotland/EDB2984/2020\|EPI_ISL_488974\|2020- | ···················· | ···················· | ····················· | ···················· | ···················· | ···················· | |
| E | 1214 | hCoV-19/Scotland/EDB3230/2020\|EPI_ISL_488975\|2020- | ···················· | ···················· | ····················· | ···················· | ···················· | ···················· | |
| E | 1215 | hCoV-19/Scotland/EDB3145/2020\|EPI_ISL_488976\|2020- | ···················· | ···················· | ····················· | ···················· | ···················· | ···················· | |
| E | 1216 | hCoV-19/Scotland/EDB2710/2020\|EPI_ISL_488977\|2020- | ···················· | ···················A | ····················· | ···················· | ···················· | ···················· | 1 |
| E | 1217 | hCoV-19/Scotland/EDB3282/2020\|EPI_ISL_488978\|2020- | ···················· | ···················A | ····················· | ···················· | ···················· | ···················· | 1 |
| E | 1218 | hCoV-19/Scotland/EDB3025/2020\|EPI_ISL_488979\|2020- | ···················· | ···················A | ····················· | ···················· | ···················· | ···················· | 1 |
| E | 1219 | hCoV-19/Scotland/EDB2701/2020\|EPI_ISL_488980\|2020- | ···················· | ···················· | ····················· | ···················· | ···················· | ···················· | |
| E | 1220 | hCoV-19/Scotland/EDB3149/2020\|EPI_ISL_488981\|2020- | ···················· | ···················· | ····················· | ···················· | ···················· | ···················· | |
| E | 1221 | hCoV-19/Scotland/EDB3013/2020\|EPI_ISL_488982\|2020- | ···················· | ···················· | ····················· | ···················· | ···················· | ···················· | |
| E | 1222 | hCoV-19/Scotland/EDB3233/2020\|EPI_ISL_488983\|2020- | ···················· | ···················· | ····················· | ···················· | ···················· | ···················· | |
| E | 1223 | hCoV-19/Scotland/EDB2790/2020\|EPI_ISL_488984\|2020- | ···················· | ···················· | ····················· | ···················· | ···················· | ···················· | |
| E | 1224 | hCoV-19/Scotland/EDB2781/2020\|EPI_ISL_488985\|2020- | ···················· | ···················· | ····················· | ···················· | ···················· | ···················· | |
| E | 1225 | hCoV-19/Scotland/EDB3084/2020\|EPI_ISL_488986\|2020- | ···················· | ···················· | ····················· | ···················· | ···················· | ···················· | |
| E | 1226 | hCoV-19/Scotland/EDB3298/2020\|EPI_ISL_488987\|2020- | ···················· | ···················A | ····················· | ···················· | ···················· | ···················· | 1 |
| E | 1227 | hCoV-19/Scotland/EDB2711/2020\|EPI_ISL_488988\|2020- | ···················· | ···················· | ····················· | ···················· | ···················· | ···················· | |

```
E 1228 hCoV-19/Scotland/EDB3221/2020|EPI_ISL_488989|2020- ···················· ···················· ····················· ···················· ···················· ····················
E 1229 hCoV-19/Scotland/EDB2974/2020|EPI_ISL_488990|2020- ···················· ···················· ····················· ···················· ···················· ····················
E 1230 hCoV-19/Scotland/EDB2947/2020|EPI_ISL_488991|2020- ···················· ···················A ····················· ···················· ···················· ···················· 1
E 1231 hCoV-19/Scotland/EDB3045/2020|EPI_ISL_488992|2020- ···················· ···················· ····················· ···················· ···················· ····················
E 1232 hCoV-19/Scotland/EDB3027/2020|EPI_ISL_488993|2020- ···················· ···················· ····················· ···················· ···················· ····················
E 1233 hCoV-19/Scotland/EDB3185/2020|EPI_ISL_488994|2020- ···················· ···················· ····················· ···················· ···················· ····················
E 1234 hCoV-19/Scotland/EDB2863/2020|EPI_ISL_488995|2020- ···················· ···················· ····················· ···················· ···················· ····················
E 1235 hCoV-19/Oman/24618/2020|EPI_ISL_491129|2020-04-17 ···················· ···················· ····················· ···················· ···················· ····················
E 1236 hCoV-19/England/BRIS-185403F/2020|EPI_ISL_492400|2 ···················· ·A·················· ····················· ···················· ···················· ····················
E 1237 hCoV-19/England/BRIS-1853FE8/2020|EPI_ISL_492401|2 ···················· ···················· ····················· ···················· ···················· ····················
E 1238 hCoV-19/England/BRIS-18532EF/2020|EPI_ISL_492403|2 ···················· ···················· ····················· ···················· ···················· ····················
E 1239 hCoV-19/England/BRIS-1853B7B/2020|EPI_ISL_492404|2 ···················· ···················· ····················· ···················· ···················· ····················
E 1240 hCoV-19/England/BRIS-185408A/2020|EPI_ISL_492405|2 ···················· ···················· ····················· ···················· ···················· ····················
E 1241 hCoV-19/England/BRIS-1853E81/2020|EPI_ISL_492406|2 ···················· ···················· ····················· ···················· ···················· ····················
E 1242 hCoV-19/England/BRIS-18531A6/2020|EPI_ISL_492407|2 ···················· ···················· ····················· ···················· ···················· ····················
E 1243 hCoV-19/England/BRIS-1853E27/2020|EPI_ISL_492410|2 ···················· ···················· ····················· ···················· ···················· ····················
E 1244 hCoV-19/England/BRIS-18543EB/2020|EPI_ISL_492411|2 ···················· ···················· ····················· ···················· ···················· ····················
E 1245 hCoV-19/England/BRIS-1853A41/2020|EPI_ISL_492412|2 ···················· ···················· ····················· ···················· ···················· ····················
E 1246 hCoV-19/England/BRIS-1853689/2020|EPI_ISL_492413|2 ···················· ·A·················· ····················· ···················· ···················· ····················
E 1247 hCoV-19/England/BRIS-185320D/2020|EPI_ISL_492415|2 ···················· ···A················ ····················· ···················· ···················· ····················
E 1248 hCoV-19/England/BRIS-1853B20/2020|EPI_ISL_492416|2 ···················· ···················· ····················· ···················· ···················· ····················
E 1249 hCoV-19/England/BRIS-1853EDC/2020|EPI_ISL_492417|2 ···················· ···················· ····················· ···················· ···················· ····················
E 1250 hCoV-19/England/BRIS-1853267/2020|EPI_ISL_492418|2 ···················· ···················· ····················· ···················· ···················· ····················
E 1251 hCoV-19/England/BRIS-1853610/2020|EPI_ISL_492420|2 ···················· ···················· ····················· ···················· ···················· ····················
E 1252 hCoV-19/England/BRIS-1854451/2020|EPI_ISL_492423|2 ···················· ···················· ····················· ···················· ···················· ····················
E 1253 hCoV-19/England/BRIS-1853B02/2020|EPI_ISL_492424|2 ···················· ···················· ····················· ···················· ···················· ····················
E 1254 hCoV-19/England/BRIS-1852FDA/2020|EPI_ISL_492425|2 ···················· ···················· ····················· ···················· ···················· ····················
E 1255 hCoV-19/England/BRIS-18536D4/2020|EPI_ISL_492426|2 ···················· ···················· ····················· ···················· ···················· ····················
E 1256 hCoV-19/England/BRIS-1854248/2020|EPI_ISL_492427|2 ···················· ···················· ····················· ···················· ···················· ····················
E 1257 hCoV-19/England/BRIS-18545B8/2020|EPI_ISL_492429|2 ···················· ···················· ····················· ···················· ···················· ····················
E 1258 hCoV-19/England/NORT-299CF7/2020|EPI_ISL_492460|20 ···················· ···················· ····················· ···················· ···················· ····················
E 1259 hCoV-19/England/SHEF-CE147/2020|EPI_ISL_493720|202 ···················· ···················· ····················· ···················· ···················· ····················
E 1260 hCoV-19/England/SHEF-CF5FF/2020|EPI_ISL_493724|202 ···················· ···················· ····················· ···················· ···················· ····················
E 1261 hCoV-19/England/SHEF-D141B/2020|EPI_ISL_493733|202 ···················· ···················· ····················· ···················· ···················· ····················
E 1262 hCoV-19/England/SHEF-D285A/2020|EPI_ISL_493735|202 ···················· ···················· ····················· ···················· ···················· ····················
E 1263 hCoV-19/England/SHEF-D33D0/2020|EPI_ISL_493739|202 ···················· ···················· ····················· ···················· ···················· ····················
E 1264 hCoV-19/Scotland/CVR111/2020|EPI_ISL_493741|2020-0 ···················· ···················· ····················· ···················· ···················· ····················
E 1265 hCoV-19/Scotland/CVR1513/2020|EPI_ISL_493742|2020- ···················· ···················· ····················· ···················· ···················· ····················
E 1266 hCoV-19/Scotland/CVR1838/2020|EPI_ISL_493744|2020- ···················· ···················· ····················· ···················· ···················· ····················
E 1276 hCoV-19/Egypt/Army-MCL001/2020|EPI_ISL_510526|2020 ···················· ···················· ····················· ···················· ···················· ····················
E 1277 hCoV-19/France/PAC-FR/2020|EPI_ISL_510527|2020-05- ···················· ···················· ····················· ···················· ···················· ····················
E 1278 hCoV-19/Bahrain/140008296/2020|EPI_ISL_510528|2020 ···················· ···················· ····················· ···················· ···················· ····················
E 1279 hCoV-19/Zambia/29/2020|EPI_ISL_510529|2020-03-16 ···················· ···················· ····················· ···················· ···················· ····················
E 1280 hCoV-19/France/PAC-BMPM/2020|EPI_ISL_510530|2020-0 ···················· ···················· ····················· ···················· ···················· ····················
E 1281 hCoV-19/Japan/PG-0470/2020|EPI_ISL_480130|2020-04- ···················· ···················· ····················· ···················· ···················· ····················
E 1282 hCoV-19/Japan/PG-0471/2020|EPI_ISL_480131|2020-04- ···················· ···················· ····················· ···················· ···················· ····················
E 1283 hCoV-19/Japan/PG-0472/2020|EPI_ISL_480132|2020-04- ···················· ···················· ····················· ···················· ···················· ····················
E 1284 hCoV-19/Japan/PG-0473/2020|EPI_ISL_480133|2020-04- ···················· ···················· ····················· ···················· ···················· ····················
E 1285 hCoV-19/Japan/PG-0476/2020|EPI_ISL_480136|2020-04- ···················· ···················· ····················· ···················· ···················· ····················
E 1286 hCoV-19/Japan/PG-0481/2020|EPI_ISL_480140|2020-04- ···················· ···················· ····················· ···················· ···················· ····················
E 1287 hCoV-19/Japan/PG-0516/2020|EPI_ISL_480174|2020-04- ···················· ···················· ····················· ···················· ···················· ····················
E 1288 hCoV-19/Japan/PG-0517/2020|EPI_ISL_480175|2020-04- ···················· ···················· ····················· ···················· ···················· ····················
E 1289 hCoV-19/Japan/PG-0518/2020|EPI_ISL_480176|2020-04- ···················· ···················· ····················· ···················· ···················· ····················
E 1290 hCoV-19/Japan/PG-0519/2020|EPI_ISL_480177|2020-04- ···················· ···················· ····················· ···················· ···················· ····················
E 1291 hCoV-19/Japan/PG-0520/2020|EPI_ISL_480178|2020-03- ···················· ···················· ····················· ···················· ···················· ····················
E 1292 hCoV-19/Japan/PG-0521/2020|EPI_ISL_480179|2020-03- ···················· ···················· ····················· ···················· ···················· ····················
E 1293 hCoV-19/Japan/PG-0522/2020|EPI_ISL_480180|2020-03- ···················· ···················· ····················· ···················· ···················· ····················
E 1294 hCoV-19/Japan/PG-0523/2020|EPI_ISL_480181|2020-03- ···················· ···················· ····················· ···················· ···················· ····················
E 1295 hCoV-19/Japan/PG-0524/2020|EPI_ISL_480182|2020-04- ···················· ···················· ····················· ···················· ···················· ····················
E 1296 hCoV-19/Japan/PG-0525/2020|EPI_ISL_480183|2020-04- ···················· ···················· ····················· ···················· ···················· ····················
E 1297 hCoV-19/Japan/PG-0526/2020|EPI_ISL_480184|2020-04- ···················· ···················· ····················· ···················· ···················· ····················
E 1298 hCoV-19/Japan/PG-0527/2020|EPI_ISL_480185|2020-04- ···················· ···················· ····················· ···················· ···················· ····················
E 1299 hCoV-19/Japan/PG-0528/2020|EPI_ISL_480186|2020-04- ···················· ···················· ····················· ···················· ···················· ····················
E 1300 hCoV-19/Japan/PG-0529/2020|EPI_ISL_480187|2020-04- ···················· ···················· ····················· ···················· ···················· ····················
E 1301 hCoV-19/Japan/PG-0530/2020|EPI_ISL_480188|2020-04- ···················· ···················· ····················· ···················· ···················· ····················
E 1302 hCoV-19/Japan/PG-0531/2020|EPI_ISL_480189|2020-04- ···················· ···················· ····················· ···················· ···················· ····················
E 1303 hCoV-19/Japan/PG-0532/2020|EPI_ISL_480190|2020-04 ···················· ···················· ····················· ···················· ···················· ····················
E 1304 hCoV-19/Japan/PG-0533/2020|EPI_ISL_480191|2020-04 ···················· ···················· ····················· ···················· ···················· ····················
E 1305 hCoV-19/Japan/PG-0534/2020|EPI_ISL_480192|2020-04 ···················· ···················· ····················· ···················· ···················· ····················
E 1306 hCoV-19/Japan/PG-0536/2020|EPI_ISL_480193|2020-04 ···················· ···················· ····················· ···················· ···················· ····················
E 1307 hCoV-19/Japan/PG-0537/2020|EPI_ISL_480194|2020-04 ···················· ···················· ····················· ···················· ···················· ····················
E 1308 hCoV-19/Japan/PG-0538/2020|EPI_ISL_480195|2020-04 ···················· ···················· ····················· ···················· ···················· ····················
E 1309 hCoV-19/Japan/PG-0541/2020|EPI_ISL_480197|2020-04- ···················· ···················· ····················· ···················· ···················· ····················
E 1310 hCoV-19/Japan/PG-0542/2020|EPI_ISL_480198|2020-03- ···················· ···················· ····················· ···················· ···················· ····················
E 1311 hCoV-19/Japan/PG-0543/2020|EPI_ISL_480199|2020-03- ···················· ···················· ····················· ···················· ···················· ····················
E 1312 hCoV-19/USA/LA-GBCL-26159751/2020|EPI_ISL_485004|2 ···················· ···················· ····················· ···················· ···················· ····················
E 1313 hCoV-19/USA/LA-GBCL-26159754/2020|EPI_ISL_485006|2 ···················· ···················· ····················· ···················· ···················· ····················
E 1314 hCoV-19/USA/LA-GBCL-26159757/2020|EPI_ISL_485008|2 ···················· ···················· ····················· ···················· ···················· ····················
E 1315 hCoV-19/USA/LA-GBCL-26159759/2020|EPI_ISL_485009|2 ···················· ···················· ····················· ···················· ···················· ····················
E 1316 hCoV-19/USA/LA-GBCL-26159764/2020|EPI_ISL_485014|2 ···················· ···················· ····················· ···················· ···················· ····················
```

| | | | | | | | | | |
|---|---|---|---|---|---|---|---|---|---|
| E | 1317 | hCoV-19/USA/LA-GBCL-26159765/2020\|EPI_ISL_485015\|2 | .................... | .................... | .................... | .................... | .................... | .................... | |
| E | 1318 | hCoV-19/USA/LA-GBCL-26159793/2020\|EPI_ISL_485018\|2 | .................... | .................... | .................... | .................... | .................... | .................... | |
| E | 1319 | hCoV-19/USA/LA-GBCL-26492434/2020\|EPI_ISL_485019\|2 | .................... | .................... | .................... | .................... | .................... | .................... | |
| E | 1320 | hCoV-19/USA/LA-GBCL-26492435/2020\|EPI_ISL_485020\|2 | .................... | .................... | .................... | .................... | .................... | .................... | |
| E | 1321 | hCoV-19/USA/LA-GBCL-26492438/2020\|EPI_ISL_485022\|2 | .................... | .................... | .................... | .................... | .................... | .................... | |
| E | 1322 | hCoV-19/USA/CA-CZB-2057/2020\|EPI_ISL_486300\|2020-0 | .................... | .................... | .................... | .................... | .................... | .................... | |
| E | 1323 | hCoV-19/USA/CA-CZB-2058/2020\|EPI_ISL_486301\|2020-0 | .................... | .................... | .................... | .................... | .................... | .................... | |
| E | 1324 | hCoV-19/USA/CA-CZB-2059/2020\|EPI_ISL_486302\|2020-0 | .................... | .................... | .................... | .................... | .................... | .................... | |
| E | 1325 | hCoV-19/USA/CA-CZB-2060/2020\|EPI_ISL_486303\|2020-0 | .................... | .................... | .................... | .................... | .................... | .................... | |
| E | 1326 | hCoV-19/USA/CA-CZB-2061/2020\|EPI_ISL_486304\|2020-0 | .................... | .................... | .................... | .................... | .................... | .................... | |
| E | 1327 | hCoV-19/USA/CA-CZB-2062/2020\|EPI_ISL_486305\|2020-0 | .................... | .................... | .................... | .................... | .................... | .................... | |
| E | 1328 | hCoV-19/USA/CA-CZB-2063/2020\|EPI_ISL_486306\|2020-0 | .................... | .................... | .................... | .................... | .................... | .................... | |
| E | 1329 | hCoV-19/USA/CA-CZB-2064/2020\|EPI_ISL_486307\|2020-0 | .................... | .................... | .................... | .................... | .................... | .................... | |
| E | 1330 | hCoV-19/USA/CA-CZB-2065/2020\|EPI_ISL_486308\|2020-0 | .................... | .................... | .................... | .................... | .................... | .................... | |
| E | 1331 | hCoV-19/USA/CA-CZB-2066/2020\|EPI_ISL_486309\|2020-0 | .................... | .................... | .................... | .................... | .................... | .................... | |
| E | 1332 | hCoV-19/USA/CA-CZB-2067/2020\|EPI_ISL_486310\|2020-0 | .................... | .................... | .................... | .................... | .................... | .................... | |
| E | 1333 | hCoV-19/USA/CA-CZB-2068/2020\|EPI_ISL_486311\|2020-0 | .................... | .................... | .................... | .................... | .................... | .................... | |
| E | 1334 | hCoV-19/USA/CA-CZB-2091/2020\|EPI_ISL_486334\|2020-0 | .................... | .................... | .................... | .................... | .................... | .................... | |
| E | 1335 | hCoV-19/USA/CA-CZB-2093/2020\|EPI_ISL_486336\|2020-0 | .................... | .................... | .................... | .................... | .................... | .................... | |
| E | 1336 | hCoV-19/USA/CA-CZB-2094/2020\|EPI_ISL_486337\|2020-0 | .................... | .................... | .................... | .................... | .................... | .................... | |
| E | 1337 | hCoV-19/USA/CA-CZB-2095/2020\|EPI_ISL_486338\|2020-0 | .................... | .................... | .................... | .................... | .................... | .................... | |
| E | 1338 | hCoV-19/USA/CA-CZB-2104/2020\|EPI_ISL_486340\|2020-0 | .................... | .................... | .................... | .................... | .................... | .................... | |
| E | 1339 | hCoV-19/USA/CA-CZB-2115/2020\|EPI_ISL_486345\|2020-0 | .................... | .................... | .................... | .................... | .................... | .................... | |
| E | 1340 | hCoV-19/USA/CA-CZB-2116/2020\|EPI_ISL_486346\|2020-0 | .................... | .................... | .................... | .................... | .................... | .................... | |
| E | 1341 | hCoV-19/USA/CA-CZB-2117/2020\|EPI_ISL_486347\|2020-0 | .................... | .................... | .................... | .................... | .................... | .................... | |
| E | 1342 | hCoV-19/USA/CA-CZB-2118/2020\|EPI_ISL_486348\|2020-0 | .................... | .................... | .................... | .................... | .................... | .................... | |
| E | 1343 | hCoV-19/USA/CA-CZB-2119/2020\|EPI_ISL_486349\|2020-0 | .................... | .................... | .................... | .................... | .................... | .................... | |
| E | 1344 | hCoV-19/USA/CA-CZB-2124/2020\|EPI_ISL_486352\|2020-0 | .................... | .................... | .................... | .................... | .................... | .................... | |
| E | 1345 | hCoV-19/USA/CA-CZB-2127/2020\|EPI_ISL_486354\|2020-0 | .................... | .................... | .................... | .................... | .................... | .................... | |
| E | 1346 | hCoV-19/USA/CA-CZB-2128/2020\|EPI_ISL_486355\|2020-0 | .................... | .................... | .................... | .................... | .................... | .................... | |
| E | 1347 | hCoV-19/England/LOND-D649E/2020\|EPI_ISL_487600\|202 | .................... | .................... | .................... | .................... | .................... | .................... | |
| E | 1348 | hCoV-19/England/BRIS-12B860/2020\|EPI_ISL_487601\|20 | .................... | .................... | .................... | .................... | .................... | .................... | |
| E | 1349 | hCoV-19/England/BRIS-12B2EA/2020\|EPI_ISL_487602\|20 | .................... | .................... | .................... | .................... | .................... | .................... | |
| E | 1350 | hCoV-19/England/BRIS-12B5D2/2020\|EPI_ISL_487604\|20 | .................... | .................... | .................... | .................... | .................... | .................... | |
| E | 1351 | hCoV-19/England/BRIS-12B7DC/2020\|EPI_ISL_487605\|20 | .................... | .................... | .................... | .................... | .................... | .................... | |
| E | 1352 | hCoV-19/England/LOND-D63CE/2020\|EPI_ISL_487606\|202 | .................... | .................... | .................... | .................... | .................... | .................... | |
| E | 1353 | hCoV-19/England/LOND-D6522/2020\|EPI_ISL_487607\|202 | .................... | .................... | .................... | .................... | .................... | .................... | |
| E | 1354 | hCoV-19/England/BRIS-12B323/2020\|EPI_ISL_487608\|20 | .................... | .................... | .................... | .................... | .................... | .................... | |
| E | 1355 | hCoV-19/England/BRIS-12B6C0/2020\|EPI_ISL_487610\|20 | .................... | .................... | .................... | .................... | .................... | .................... | |
| E | 1356 | hCoV-19/England/BRIS-12B3D8/2020\|EPI_ISL_487611\|20 | .................... | .................... | .................... | .................... | .................... | .................... | |
| E | 1357 | hCoV-19/England/BRIS-12B6B1/2020\|EPI_ISL_487612\|20 | .................... | .................... | .................... | .................... | .................... | .................... | |
| E | 1358 | hCoV-19/England/BRIS-12B6FD/2020\|EPI_ISL_487613\|20 | .................... | .................... | .................... | .................... | .................... | .................... | |
| E | 1359 | hCoV-19/England/LOND-D64BC/2020\|EPI_ISL_487615\|202 | .................... | .................... | .................... | .................... | .................... | .................... | |
| E | 1360 | hCoV-19/England/LOND-D6504/2020\|EPI_ISL_487616\|202 | .................... | .................... | .................... | .................... | .................... | .................... | |
| E | 1361 | hCoV-19/England/BRIS-12B45D/2020\|EPI_ISL_487617\|20 | .................... | .................... | .................... | .................... | .................... | .................... | |
| E | 1362 | hCoV-19/England/BRIS-12B499/2020\|EPI_ISL_487618\|20 | .................... | .................... | .................... | .................... | .................... | .................... | |
| E | 1363 | hCoV-19/England/BRIS-12B587/2020\|EPI_ISL_487619\|20 | .................... | .................... | .................... | .................... | .................... | .................... | |
| E | 1364 | hCoV-19/England/BRIS-12B2DB/2020\|EPI_ISL_487620\|20 | .................... | .................... | .................... | .................... | .................... | .................... | |
| E | 1365 | hCoV-19/England/BRIS-12B4C6/2020\|EPI_ISL_487622\|20 | .................... | .................... | .................... | .................... | .................... | .................... | |
| E | 1366 | hCoV-19/England/BRIS-12B6EE/2020\|EPI_ISL_487623\|20 | .................... | .................... | .................... | .................... | .................... | .................... | |
| E | 1367 | hCoV-19/England/BRIS-12B402/2020\|EPI_ISL_487624\|20 | .................... | .................... | .................... | .................... | .................... | .................... | |
| E | 1368 | hCoV-19/England/LOND-D6689/2020\|EPI_ISL_487625\|202 | .................... | .................... | .................... | .................... | .................... | .................... | |
| E | 1369 | hCoV-19/England/LOND-D6610/2020\|EPI_ISL_487626\|202 | .................... | .................... | .................... | .................... | .................... | .................... | |
| E | 1370 | hCoV-19/England/LOND-D658C/2020\|EPI_ISL_487627\|202 | .................... | .................... | .................... | .................... | .................... | .................... | |
| E | 1371 | hCoV-19/England/LOND-D6601/2020\|EPI_ISL_487628\|202 | .................... | .................... | .................... | .................... | .................... | .................... | |
| E | 1372 | hCoV-19/England/BRIS-12B87F/2020\|EPI_ISL_487629\|20 | .................... | .................... | .................... | .................... | .................... | .................... | |
| E | 1373 | hCoV-19/England/LOND-D6434/2020\|EPI_ISL_487630\|202 | .................... | .................... | .................... | .................... | .................... | .................... | |
| E | 1374 | hCoV-19/England/LOND-D667A/2020\|EPI_ISL_487631\|202 | .................... | .................... | .................... | .................... | .................... | .................... | |
| E | 1375 | hCoV-19/England/LOND-D66B6/2020\|EPI_ISL_487632\|202 | .................... | .................... | .................... | .................... | .................... | .................... | |
| E | 1376 | hCoV-19/England/LOND-D666B/2020\|EPI_ISL_487633\|202 | .................... | .................... | .................... | .................... | .................... | .................... | |
| E | 1377 | hCoV-19/England/BRIS-12B693/2020\|EPI_ISL_487634\|20 | .................... | .................... | .................... | .................... | .................... | .................... | |
| E | 1378 | hCoV-19/England/LOND-D962C/2020\|EPI_ISL_487635\|202 | .................... | .................... | .................... | .................... | .................... | .................... | |
| E | 1379 | hCoV-19/England/LOND-D9783/2020\|EPI_ISL_487636\|202 | .................... | .................... | .................... | .................... | .................... | .................... | |
| E | 1380 | hCoV-19/England/LOND-D9808/2020\|EPI_ISL_487637\|202 | .................... | .................... | .................... | .................... | .................... | .................... | |
| E | 1381 | hCoV-19/England/LOND-D96FF/2020\|EPI_ISL_487638\|202 | .................... | .................... | .................... | .................... | .................... | .................... | |
| E | 1382 | hCoV-19/England/LOND-D970B/2020\|EPI_ISL_487639\|202 | .................... | .................... | .................... | .................... | .................... | .................... | |
| E | 1383 | hCoV-19/England/LOND-D963B/2020\|EPI_ISL_487641\|202 | .................... | .................... | .................... | .................... | .................... | .................... | |
| E | 1384 | hCoV-19/Scotland/EDB5977/2020\|EPI_ISL_487665\|2020- | .................... | .................... | .................... | .................... | .................... | .................... | |
| E | 1385 | hCoV-19/Scotland/EDB5499/2020\|EPI_ISL_487666\|2020- | .................... | .................... | .................... | .................... | .................... | .................... | |
| E | 1386 | hCoV-19/Scotland/EDB6038/2020\|EPI_ISL_487667\|2020- | .................... | .A.................. | .................... | .................... | .................... | .................... | |
| E | 1387 | hCoV-19/Scotland/EDB5849/2020\|EPI_ISL_487668\|2020- | .................... | .A.................. | .................... | .................... | .................... | .................... | |
| E | 1388 | hCoV-19/Scotland/EDB6437/2020\|EPI_ISL_487669\|2020- | .................... | .................... | .................... | .................... | .................... | .................... | |
| E | 1389 | hCoV-19/Scotland/EDB3386/2020\|EPI_ISL_487670\|2020- | .................... | .A.................. | .................... | .................... | .................... | .................... | |
| E | 1390 | hCoV-19/Scotland/EDB5509/2020\|EPI_ISL_487676\|2020- | .................... | .................... | .................... | .................... | .................... | .................... | |
| E | 1391 | hCoV-19/Scotland/EDB4208/2020\|EPI_ISL_487677\|2020- | .................... | ...................A | .................... | .................... | .................... | .................... | 1 |
| E | 1392 | hCoV-19/Scotland/EDB2776/2020\|EPI_ISL_487678\|2020- | .................... | .................... | .................... | .................... | .................... | .................... | |
| E | 1393 | hCoV-19/Scotland/EDB3511/2020\|EPI_ISL_487679\|2020- | .................... | .................... | .................... | .................... | .................... | .................... | |
| E | 1394 | hCoV-19/Scotland/EDB5496/2020\|EPI_ISL_487680\|2020- | .................... | .................... | .................... | .................... | .................... | .................... | |
| E | 1395 | hCoV-19/Scotland/EDB3505/2020\|EPI_ISL_487681\|2020- | .................... | ...................A | .................... | .................... | .................... | .................... | 1 |
| E | 1396 | hCoV-19/Scotland/EDB6044/2020\|EPI_ISL_487682\|2020- | .................... | ...................A | .................... | .................... | .................... | .................... | 1 |

| | | | | | | | | | |
|---|---|---|---|---|---|---|---|---|---|
| E | 1397 | hCoV-19/Scotland/EDB5960/2020\|EPI_ISL_487683\|2020- | .................... | .................... | .................... | .................... | .................... | .................... | |
| E | 1398 | hCoV-19/Scotland/EDB2823/2020\|EPI_ISL_487684\|2020- | .................... | .................... | .................... | .................... | .................... | .................... | |
| E | 1399 | hCoV-19/Scotland/EDB5641/2020\|EPI_ISL_487685\|2020- | .................... | .................... | .................... | .................... | .................... | .................... | |
| E | 1400 | hCoV-19/Scotland/EDB3419/2020\|EPI_ISL_487686\|2020- | .................... | ...................A | .................... | .................... | .................... | .................... | 1 |
| E | 1401 | hCoV-19/Scotland/EDB2697/2020\|EPI_ISL_488900\|2020- | .................... | .A.................. | .................... | .................... | .................... | .................... | |
| E | 1402 | hCoV-19/Scotland/EDB2707/2020\|EPI_ISL_488901\|2020- | .................... | ...................A | .................... | .................... | .................... | .................... | 1 |
| E | 1403 | hCoV-19/Scotland/EDB2874/2020\|EPI_ISL_488902\|2020- | .................... | ...................A | .................... | .................... | .................... | .................... | 1 |
| E | 1404 | hCoV-19/Scotland/EDB2960/2020\|EPI_ISL_488903\|2020- | .................... | ...................A | .................... | .................... | .................... | .................... | 1 |
| E | 1405 | hCoV-19/Scotland/EDB3235/2020\|EPI_ISL_488904\|2020- | .................... | .................... | .................... | .................... | .................... | .................... | |
| E | 1406 | hCoV-19/Scotland/EDB2767/2020\|EPI_ISL_488905\|2020- | .................... | .................... | .................... | .................... | .................... | .................... | |
| E | 1407 | hCoV-19/Scotland/EDB2687/2020\|EPI_ISL_488906\|2020- | .................... | .................... | .................... | .................... | .................... | .................... | |
| E | 1408 | hCoV-19/Scotland/EDB3260/2020\|EPI_ISL_488907\|2020- | .................... | .................... | .................... | .................... | .................... | .................... | |
| E | 1409 | hCoV-19/Scotland/EDB3227/2020\|EPI_ISL_488908\|2020- | .................... | ...................A | .................... | .................... | .................... | .................... | 1 |
| E | 1410 | hCoV-19/Scotland/EDB2846/2020\|EPI_ISL_488909\|2020- | .................... | .................... | .................... | .................... | .................... | .................... | |
| E | 1411 | hCoV-19/Scotland/EDB3050/2020\|EPI_ISL_488910\|2020- | .................... | .................... | .................... | .................... | .................... | .................... | |
| E | 1412 | hCoV-19/Scotland/EDB2968/2020\|EPI_ISL_488911\|2020- | .................... | .................... | .................... | .................... | .................... | .................... | |
| E | 1413 | hCoV-19/Scotland/EDB2773/2020\|EPI_ISL_488912\|2020- | .................... | ...................A | .................... | .................... | .................... | .................... | 1 |
| E | 1414 | hCoV-19/Scotland/EDB3073/2020\|EPI_ISL_488913\|2020- | .................... | ...................A | .................... | .................... | .................... | .................... | 1 |
| E | 1415 | hCoV-19/Scotland/EDB3123/2020\|EPI_ISL_488914\|2020- | .................... | .................... | .................... | .................... | .................... | .................... | |
| E | 1416 | hCoV-19/Scotland/EDB2904/2020\|EPI_ISL_488915\|2020- | .................... | .................... | .................... | .................... | .................... | .................... | |
| E | 1417 | hCoV-19/Scotland/EDB3093/2020\|EPI_ISL_488916\|2020- | .................... | ...................A | .................... | .................... | .................... | .................... | 1 |
| E | 1418 | hCoV-19/Scotland/EDB3234/2020\|EPI_ISL_488917\|2020- | .................... | .................... | .................... | .................... | .................... | .................... | |
| E | 1419 | hCoV-19/Scotland/EDB2819/2020\|EPI_ISL_488918\|2020- | .................... | .................... | .................... | .................... | .................... | .................... | |
| E | 1420 | hCoV-19/Scotland/EDB3188/2020\|EPI_ISL_488919\|2020- | .................... | ...................A | .................... | .................... | .................... | .................... | 1 |
| E | 1421 | hCoV-19/Scotland/EDB3042/2020\|EPI_ISL_488920\|2020- | .................... | .................... | .................... | .................... | .................... | .................... | |
| E | 1422 | hCoV-19/Scotland/EDB3271/2020\|EPI_ISL_488921\|2020- | .................... | ...................A | .................... | .................... | .................... | .................... | 1 |
| E | 1423 | hCoV-19/Scotland/EDB3183/2020\|EPI_ISL_488922\|2020- | .................... | ...................A | .................... | .................... | .................... | .................... | 1 |
| E | 1424 | hCoV-19/Scotland/EDB3182/2020\|EPI_ISL_488923\|2020- | .................... | .................... | .................... | .................... | .................... | .................... | |
| E | 1425 | hCoV-19/Scotland/EDB2733/2020\|EPI_ISL_488924\|2020- | .................... | ...................A | .................... | .................... | .................... | .................... | 1 |
| E | 1426 | hCoV-19/Scotland/EDB2754/2020\|EPI_ISL_488925\|2020- | .................... | .................... | .................... | .................... | .................... | .................... | |
| E | 1427 | hCoV-19/Scotland/EDB2799/2020\|EPI_ISL_488926\|2020- | .................... | .................... | .................... | .................... | .................... | .................... | |
| E | 1428 | hCoV-19/Scotland/EDB3075/2020\|EPI_ISL_488927\|2020- | .................... | ...................A | .................... | .................... | .................... | .................... | 1 |
| E | 1429 | hCoV-19/Scotland/EDB3280/2020\|EPI_ISL_488928\|2020- | .................... | ...................A | .................... | .................... | .................... | .................... | 1 |
| E | 1430 | hCoV-19/Scotland/EDB3043/2020\|EPI_ISL_488929\|2020- | .................... | .................... | .................... | .................... | .................... | .................... | |
| E | 1431 | hCoV-19/Scotland/EDB2885/2020\|EPI_ISL_488930\|2020- | .................... | ...................A | .................... | .................... | .................... | .................... | 1 |
| E | 1432 | hCoV-19/Scotland/EDB2869/2020\|EPI_ISL_488931\|2020- | .................... | .................... | .................... | .................... | .................... | .................... | |
| E | 1433 | hCoV-19/Scotland/EDB3031/2020\|EPI_ISL_488932\|2020- | .................... | ...................A | .................... | .................... | .................... | .................... | 1 |
| E | 1434 | hCoV-19/Scotland/EDB3214/2020\|EPI_ISL_488933\|2020- | .................... | .................... | .................... | .................... | .................... | .................... | |
| E | 1435 | hCoV-19/Scotland/EDB3213/2020\|EPI_ISL_488934\|2020- | .................... | .................... | .................... | .................... | .................... | .................... | |
| E | 1436 | hCoV-19/Scotland/EDB3186/2020\|EPI_ISL_488935\|2020- | .................... | ...................A | .................... | .................... | .................... | .................... | 1 |
| E | 1437 | hCoV-19/Scotland/EDB3232/2020\|EPI_ISL_488936\|2020- | .................... | .................... | .................... | .................... | .................... | .................... | |
| E | 1438 | hCoV-19/Scotland/EDB2768/2020\|EPI_ISL_488937\|2020- | .................... | .................... | .................... | .................... | .................... | .................... | |
| E | 1439 | hCoV-19/Scotland/EDB3002/2020\|EPI_ISL_488938\|2020- | .................... | ...................A | .................... | .................... | .................... | .................... | 1 |
| E | 1440 | hCoV-19/Scotland/EDB2955/2020\|EPI_ISL_488939\|2020- | .................... | ...................A | .................... | .................... | .................... | .................... | 1 |
| E | 1441 | hCoV-19/Scotland/EDB2877/2020\|EPI_ISL_488940\|2020- | .................... | ...................A | .................... | .................... | .................... | .................... | 1 |
| E | 1442 | hCoV-19/Scotland/EDB2979/2020\|EPI_ISL_488942\|2020- | .................... | ...................A | .................... | .................... | .................... | .................... | 1 |
| E | 1443 | hCoV-19/Scotland/EDB2693/2020\|EPI_ISL_488943\|2020- | .................... | .................... | .................... | .................... | .................... | .................... | |
| E | 1444 | hCoV-19/Scotland/EDB3313/2020\|EPI_ISL_488944\|2020- | .................... | ...................A | .................... | .................... | .................... | .................... | 1 |
| E | 1445 | hCoV-19/Scotland/EDB2908/2020\|EPI_ISL_488945\|2020- | .................... | ...................A | .................... | .................... | .................... | .................... | 1 |
| E | 1446 | hCoV-19/Scotland/EDB3270/2020\|EPI_ISL_488946\|2020- | .................... | ...................A | .................... | .................... | .................... | .................... | 1 |
| E | 1447 | hCoV-19/Scotland/EDB2805/2020\|EPI_ISL_488947\|2020- | .................... | .................... | .................... | .................... | .................... | .................... | |
| E | 1448 | hCoV-19/Scotland/EDB3024/2020\|EPI_ISL_488948\|2020- | .................... | .................... | .................... | .................... | .................... | .................... | |
| E | 1449 | hCoV-19/Scotland/EDB3014/2020\|EPI_ISL_488949\|2020- | .................... | .................... | .................... | .................... | .................... | .................... | |
| E | 1450 | hCoV-19/Scotland/EDB3296/2020\|EPI_ISL_488950\|2020- | .................... | ...................A | .................... | .................... | .................... | .................... | 1 |
| E | 1451 | hCoV-19/Scotland/EDB3110/2020\|EPI_ISL_488951\|2020- | .................... | .................... | .................... | .................... | .................... | .................... | |
| E | 1452 | hCoV-19/Scotland/EDB3231/2020\|EPI_ISL_488952\|2020- | .................... | .................... | .................... | .................... | .................... | .................... | |
| E | 1453 | hCoV-19/Scotland/EDB3287/2020\|EPI_ISL_488953\|2020- | .................... | .................... | .................... | .................... | .................... | .................... | |
| E | 1454 | hCoV-19/Scotland/EDB3098/2020\|EPI_ISL_488954\|2020- | .................... | .................... | .................... | .................... | .................... | .................... | |
| E | 1455 | hCoV-19/Scotland/EDB2792/2020\|EPI_ISL_488955\|2020- | .................... | ...................A | .................... | .................... | .................... | .................... | 1 |
| E | 1456 | hCoV-19/Scotland/EDB3037/2020\|EPI_ISL_488956\|2020- | .................... | ...................A | .................... | .................... | .................... | .................... | 1 |
| E | 1457 | hCoV-19/Scotland/EDB2902/2020\|EPI_ISL_488957\|2020- | .................... | .................... | .................... | .................... | .................... | .................... | |
| E | 1458 | hCoV-19/Scotland/EDB2734/2020\|EPI_ISL_488958\|2020- | .................... | .................... | .................... | .................... | .................... | .................... | |
| E | 1459 | hCoV-19/Scotland/EDB3181/2020\|EPI_ISL_488959\|2020- | .................... | .................... | .................... | .................... | .................... | .................... | |
| E | 1460 | hCoV-19/Scotland/EDB3236/2020\|EPI_ISL_488960\|2020- | .................... | ...................A | .................... | .................... | .................... | .................... | 1 |
| E | 1461 | hCoV-19/Scotland/EDB2843/2020\|EPI_ISL_488961\|2020- | .................... | .................... | .................... | .................... | .................... | .................... | |
| E | 1462 | hCoV-19/Scotland/EDB3196/2020\|EPI_ISL_488962\|2020- | .................... | .................... | .................... | .................... | .................... | .................... | |
| E | 1463 | hCoV-19/Scotland/EDB3158/2020\|EPI_ISL_488963\|2020- | .................... | .................... | .................... | .................... | .................... | .................... | |
| E | 1464 | hCoV-19/Scotland/EDB2793/2020\|EPI_ISL_488964\|2020- | .................... | ...................A | .................... | .................... | .................... | .................... | 1 |
| E | 1465 | hCoV-19/Scotland/EDB2853/2020\|EPI_ISL_488965\|2020- | .................... | .................... | .................... | .................... | .................... | .................... | |
| E | 1466 | hCoV-19/Scotland/EDB3278/2020\|EPI_ISL_488966\|2020- | .................... | .................... | .................... | .................... | .................... | .................... | |
| E | 1467 | hCoV-19/Scotland/EDB3162/2020\|EPI_ISL_488967\|2020- | .................... | .................... | .................... | .................... | .................... | .................... | |
| E | 1468 | hCoV-19/Scotland/EDB3243/2020\|EPI_ISL_488968\|2020- | .................... | .................... | .................... | .................... | .................... | .................... | |
| E | 1469 | hCoV-19/Scotland/EDB2704/2020\|EPI_ISL_488969\|2020- | .................... | .................... | .................... | .................... | .................... | .................... | |
| E | 1470 | hCoV-19/Scotland/EDB2828/2020\|EPI_ISL_488970\|2020- | .................... | .................... | .................... | .................... | .................... | .................... | |
| E | 1471 | hCoV-19/Scotland/EDB3286/2020\|EPI_ISL_488971\|2020- | .................... | .................... | .................... | .................... | .................... | .................... | |
| E | 1472 | hCoV-19/Scotland/EDB3135/2020\|EPI_ISL_488972\|2020- | .................... | .................... | .................... | .................... | .................... | .................... | |
| E | 1473 | hCoV-19/Scotland/EDB2896/2020\|EPI_ISL_488973\|2020- | .................... | .............A...... | .................... | .................... | .................... | .................... | |
| E | 1474 | hCoV-19/Scotland/EDB2731/2020\|EPI_ISL_488996\|2020- | .................... | .................... | .................... | .................... | .................... | .................... | |
| E | 1475 | hCoV-19/Scotland/EDB2787/2020\|EPI_ISL_488997\|2020- | .................... | ...................A | .................... | .................... | .................... | .................... | 1 |
| E | 1476 | hCoV-19/Scotland/EDB3245/2020\|EPI_ISL_488998\|2020- | .................... | .................... | .................... | .................... | .................... | .................... | |

| | | | | | | | | |
|---|---|---|---|---|---|---|---|---|
| E | 1477 | hCoV-19/Scotland/EDB2950/2020\|EPI_ISL_488999\|2020- | .................... | .................... | .................... | .................... | .................... | .................... |
| E | 1478 | hCoV-19/Australia/SAP181/2020\|EPI_ISL_492122\|2020- | .................... | .................... | .................... | .................... | .................... | .................... |
| E | 1479 | hCoV-19/Australia/SAP183/2020\|EPI_ISL_492123\|2020- | .................... | .................... | .................... | .................... | .................... | .................... |
| E | 1480 | hCoV-19/Australia/SAP185/2020\|EPI_ISL_492124\|2020- | .................... | .................... | .................... | .................... | .................... | .................... |
| E | 1481 | hCoV-19/Australia/SAP186/2020\|EPI_ISL_492125\|2020- | .................... | .................... | .................... | .................... | .................... | .................... |
| E | 1482 | hCoV-19/Australia/SAP187/2020\|EPI_ISL_492126\|2020- | .................... | .................... | .................... | .................... | .................... | .................... |
| E | 1483 | hCoV-19/Australia/SAP190/2020\|EPI_ISL_492127\|2020- | .................... | .................... | .................... | .................... | .................... | .................... |
| E | 1484 | hCoV-19/Australia/SAP191/2020\|EPI_ISL_492128\|2020- | .................... | .................... | .................... | .................... | .................... | .................... |
| E | 1485 | hCoV-19/Australia/SAP194/2020\|EPI_ISL_492129\|2020- | .................... | .................... | .................... | .................... | .................... | .................... |
| E | 1486 | hCoV-19/Australia/SAP388/2020\|EPI_ISL_492153\|2020- | .................... | .................... | .................... | .................... | .................... | .................... |
| E | 1487 | hCoV-19/Australia/SAP389/2020\|EPI_ISL_492154\|2020- | .................... | .................... | .................... | .................... | .................... | .................... |
| E | 1488 | hCoV-19/Australia/SAP390/2020\|EPI_ISL_492155\|2020- | .................... | .................... | .................... | .................... | .................... | .................... |
| E | 1489 | hCoV-19/Australia/SAP391/2020\|EPI_ISL_492156\|2020- | .................... | .................... | .................... | .................... | .................... | .................... |
| E | 1490 | hCoV-19/Australia/SAP392/2020\|EPI_ISL_492157\|2020- | .................... | .................... | .................... | .................... | .................... | .................... |
| E | 1491 | hCoV-19/Australia/SAP393/2020\|EPI_ISL_492158\|2020- | .................... | .................... | .................... | .................... | .................... | .................... |
| E | 1492 | hCoV-19/Australia/SAP394/2020\|EPI_ISL_492159\|2020- | .................... | .................... | .................... | .................... | .................... | .................... |
| E | 1493 | hCoV-19/Australia/SAP395/2020\|EPI_ISL_492160\|2020- | .................... | .................... | .................... | .................... | .................... | .................... |
| E | 1494 | hCoV-19/Australia/SAP396/2020\|EPI_ISL_492161\|2020- | .................... | .................... | .................... | .................... | .................... | .................... |
| E | 1495 | hCoV-19/Australia/SAP397/2020\|EPI_ISL_492162\|2020- | .................... | .................... | .................... | .................... | .................... | .................... |
| E | 1496 | hCoV-19/Australia/SAP398/2020\|EPI_ISL_492163\|2020- | .................... | .................... | .................... | .................... | .................... | .................... |
| E | 1497 | hCoV-19/Australia/SAP399/2020\|EPI_ISL_492164\|2020- | .................... | .................... | .................... | .................... | .................... | .................... |
| E | 1498 | hCoV-19/Australia/SAP400/2020\|EPI_ISL_492165\|2020- | .................... | .................... | .................... | .................... | .................... | .................... |
| E | 1499 | hCoV-19/Australia/SAP401/2020\|EPI_ISL_492166\|2020- | .................... | .................... | .................... | .................... | .................... | .................... |
| E | 1500 | hCoV-19/USA/AR-UAMS001/2020\|EPI_ISL_492181\|2020-07 | .................... | .................... | .................... | .................... | .................... | .................... |
| E | 1501 | hCoV-19/USA/AR-UAMS002/2020\|EPI_ISL_492182\|2020-07 | .................... | .................... | .................... | .................... | .................... | .................... |
| E | 1502 | hCoV-19/Italy/CAM-INTPas-1/2020\|EPI_ISL_492184\|202 | .................... | .................... | .................... | .................... | .................... | .................... |
| E | 1503 | hCoV-19/England/BRIS-13273C/2020\|EPI_ISL_492185\|20 | .................... | .................... | .................... | .................... | .................... | .................... |
| E | 1504 | hCoV-19/England/BRIS-132839/2020\|EPI_ISL_492186\|20 | .................... | .................... | .................... | .................... | .................... | .................... |
| E | 1505 | hCoV-19/England/BRIS-132848/2020\|EPI_ISL_492187\|20 | .................... | .................... | .................... | .................... | .................... | .................... |
| E | 1506 | hCoV-19/England/CAMB-1B451A/2020\|EPI_ISL_492190\|20 | .................... | .................... | .................... | .................... | .................... | .................... |
| E | 1507 | hCoV-19/England/CAMB-1B45DE/2020\|EPI_ISL_492191\|20 | .................... | .................... | .................... | .................... | .................... | .................... |
| E | 1508 | hCoV-19/England/CAMB-1B488A/2020\|EPI_ISL_492192\|20 | .................... | .................... | .................... | .................... | .................... | .................... |
| E | 1509 | hCoV-19/England/CAMB-1B4820/2020\|EPI_ISL_492193\|20 | .................... | .................... | .................... | .................... | .................... | .................... |
| E | 1510 | hCoV-19/England/CAMB-1B4565/2020\|EPI_ISL_492194\|20 | .................... | .................... | .................... | .................... | .................... | .................... |
| E | 1511 | hCoV-19/England/CAMB-1B4626/2020\|EPI_ISL_492196\|20 | .................... | .................... | .................... | .................... | .................... | .................... |
| E | 1512 | hCoV-19/England/BRIS-132602/2020\|EPI_ISL_492197\|20 | .................... | .................... | .................... | .................... | .................... | .................... |
| E | 1513 | hCoV-19/England/CAMB-1B396A/2020\|EPI_ISL_492198\|20 | .................... | .................... | .................... | .................... | .................... | .................... |
| E | 1514 | hCoV-19/England/CAMB-1B39F1/2020\|EPI_ISL_492199\|20 | .................... | .................... | .................... | .................... | .................... | .................... |
| E | 1515 | hCoV-19/England/BIRM-61717/2020\|EPI_ISL_493451\|202 | .................... | .................... | .................... | .................... | .................... | .................... |
| E | 1516 | hCoV-19/England/NORT-285A12/2020\|EPI_ISL_493452\|20 | .................... | .................... | .................... | .................... | .................... | .................... |
| E | 1517 | hCoV-19/England/NORT-285A5E/2020\|EPI_ISL_493453\|20 | .................... | .................... | .................... | .................... | .................... | .................... |
| E | 1518 | hCoV-19/England/NORT-285AB8/2020\|EPI_ISL_493454\|20 | .................... | .................... | .................... | .................... | .................... | .................... |
| E | 1519 | hCoV-19/England/NORT-285AF4/2020\|EPI_ISL_493455\|20 | .................... | .................... | .................... | .................... | .................... | .................... |
| E | 1520 | hCoV-19/England/NORT-285B79/2020\|EPI_ISL_493456\|20 | .................... | .................... | .................... | .................... | .................... | .................... |
| E | 1521 | hCoV-19/England/NORT-285C49/2020\|EPI_ISL_493458\|20 | .................... | .................... | .................... | .................... | .................... | .................... |
| E | 1522 | hCoV-19/England/NORT-285C58/2020\|EPI_ISL_493459\|20 | .................... | .................... | .................... | .................... | .................... | .................... |
| E | 1523 | hCoV-19/England/NORT-2991DA/2020\|EPI_ISL_493484\|20 | .................... | .................... | .................... | .................... | .................... | .................... |
| E | 1524 | hCoV-19/England/NORT-299398/2020\|EPI_ISL_493485\|20 | .................... | .................... | .................... | .................... | .................... | .................... |
| E | 1525 | hCoV-19/England/NORT-2993A7/2020\|EPI_ISL_493486\|20 | .................... | .................... | .................... | .................... | .................... | .................... |
| E | 1526 | hCoV-19/England/NORT-2993E3/2020\|EPI_ISL_493487\|20 | .................... | .................... | .................... | .................... | .................... | .................... |
| E | 1527 | hCoV-19/England/NORT-29AF0C/2020\|EPI_ISL_493488\|20 | .................... | .................... | .................... | .................... | .................... | .................... |
| E | 1528 | hCoV-19/England/NORT-29AFFD/2020\|EPI_ISL_493489\|20 | .................... | .................... | .................... | .................... | .................... | .................... |
| E | 1529 | hCoV-19/England/NORT-29B017/2020\|EPI_ISL_493490\|20 | .................... | .................... | .................... | .................... | .................... | .................... |
| E | 1530 | hCoV-19/England/NORT-29B1BA/2020\|EPI_ISL_493491\|20 | .................... | .................... | .................... | .................... | .................... | .................... |
| E | 1531 | hCoV-19/England/NORT-29B387/2020\|EPI_ISL_493492\|20 | .................... | .................... | .................... | .................... | .................... | .................... |
| E | 1532 | hCoV-19/England/NORT-29B42A/2020\|EPI_ISL_493493\|20 | .................... | .................... | .................... | .................... | .................... | .................... |
| E | 1533 | hCoV-19/England/NORT-29B642/2020\|EPI_ISL_493496\|20 | .................... | .................... | .................... | .................... | .................... | .................... |
| E | 1534 | hCoV-19/England/NORT-29B660/2020\|EPI_ISL_493497\|20 | .................... | .................... | .................... | .................... | .................... | .................... |
| E | 1535 | hCoV-19/England/NORT-29DD2D/2020\|EPI_ISL_493498\|20 | .................... | .................... | .................... | .................... | .................... | .................... |
| E | 1536 | hCoV-19/England/NORT-29DE1B/2020\|EPI_ISL_493499\|20 | .................... | .................... | .................... | .................... | .................... | .................... |
| E | 1537 | hCoV-19/Japan/PG-0434/2020\|EPI_ISL_480100\|2020-04- | .................... | .................... | .................... | .................... | .................... | .................... |
| E | 1538 | hCoV-19/Japan/PG-0435/2020\|EPI_ISL_480101\|2020-04- | .................... | .................... | .................... | .................... | .................... | .................... |
| E | 1539 | hCoV-19/Japan/PG-0436/2020\|EPI_ISL_480102\|2020-04- | .................... | .................... | .................... | .................... | .................... | .................... |
| E | 1540 | hCoV-19/Japan/PG-0437/2020\|EPI_ISL_480103\|2020-03- | .................... | .................... | .................... | .................... | .................... | .................... |
| E | 1541 | hCoV-19/Japan/PG-0439/2020\|EPI_ISL_480104\|2020-03- | .................... | .................... | .................... | .................... | .................... | .................... |
| E | 1542 | hCoV-19/Japan/PG-0440/2020\|EPI_ISL_480105\|2020-03- | .................... | .................... | .................... | .................... | .................... | .................... |
| E | 1543 | hCoV-19/Japan/PG-0441/2020\|EPI_ISL_480106\|2020-03- | .................... | .................... | .................... | .................... | .................... | .................... |
| E | 1544 | hCoV-19/Japan/PG-0442/2020\|EPI_ISL_480107\|2020-03- | .................... | .................... | .................... | .................... | .................... | .................... |
| E | 1545 | hCoV-19/Japan/PG-0443/2020\|EPI_ISL_480108\|2020-03- | .................... | .................... | .................... | .................... | .................... | .................... |
| E | 1546 | hCoV-19/Japan/PG-0445/2020\|EPI_ISL_480109\|2020-03- | .................... | .................... | .................... | .................... | .................... | .................... |
| E | 1547 | hCoV-19/Japan/PG-0446/2020\|EPI_ISL_480110\|2020-03- | .................... | .................... | .................... | .................... | .................... | .................... |
| E | 1548 | hCoV-19/Japan/PG-0448/2020\|EPI_ISL_480111\|2020-03- | .................... | .................... | .................... | .................... | .................... | .................... |
| E | 1549 | hCoV-19/Japan/PG-0449/2020\|EPI_ISL_480112\|2020-03- | .................... | .................... | .................... | .................... | .................... | .................... |
| E | 1550 | hCoV-19/Japan/PG-0450/2020\|EPI_ISL_480113\|2020-03- | .................... | .................... | .................... | .................... | .................... | .................... |
| E | 1551 | hCoV-19/Japan/PG-0452/2020\|EPI_ISL_480114\|2020-03- | .................... | .................... | .................... | .................... | .................... | .................... |
| E | 1552 | hCoV-19/Japan/PG-0453/2020\|EPI_ISL_480115\|2020-03- | .................... | .................... | .................... | .................... | .................... | .................... |
| E | 1553 | hCoV-19/Japan/PG-0454/2020\|EPI_ISL_480116\|2020-03- | .................... | .................... | .................... | .................... | .................... | .................... |
| E | 1554 | hCoV-19/Japan/PG-0455/2020\|EPI_ISL_480117\|2020-03- | .................... | .................... | .................... | .................... | .................... | .................... |
| E | 1555 | hCoV-19/Japan/PG-0457/2020\|EPI_ISL_480118\|2020-03- | .................... | .................... | .................... | .................... | .................... | .................... |
| E | 1556 | hCoV-19/Japan/PG-0458/2020\|EPI_ISL_480119\|2020-03- | .................... | .................... | .................... | .................... | .................... | .................... |

| | | | | | | | |
|---|---|---|---|---|---|---|---|
| E | 1557 hCoV-19/Japan/PG-0459/2020\|EPI_ISL_480120\|2020-03- | ···················· | ···················· | ···················· | ···················· | ···················· | ···················· |
| E | 1558 hCoV-19/Japan/PG-0460/2020\|EPI_ISL_480121\|2020-03- | ···················· | ···················· | ···················· | ···················· | ···················· | ···················· |
| E | 1559 hCoV-19/Japan/PG-0461/2020\|EPI_ISL_480122\|2020-03- | ···················· | ···················· | ···················· | ···················· | ···················· | ···················· |
| E | 1560 hCoV-19/Japan/PG-0462/2020\|EPI_ISL_480123\|2020-03- | ···················· | ···················· | ···················· | ···················· | ···················· | ···················· |
| E | 1561 hCoV-19/Japan/PG-0463/2020\|EPI_ISL_480124\|2020-03- | ···················· | ···················· | ···················· | ···················· | ···················· | ···················· |
| E | 1562 hCoV-19/Japan/PG-0464/2020\|EPI_ISL_480125\|2020-03- | ···················· | ···················· | ···················· | ···················· | ···················· | ···················· |
| E | 1563 hCoV-19/Japan/PG-0465/2020\|EPI_ISL_480126\|2020-04- | ···················· | ···················· | ···················· | ···················· | ···················· | ···················· |
| E | 1564 hCoV-19/Japan/PG-0466/2020\|EPI_ISL_480127\|2020-04- | ···················· | ···················· | ···················· | ···················· | ···················· | ···················· |
| E | 1565 hCoV-19/Japan/PG-0468/2020\|EPI_ISL_480128\|2020-03- | ···················· | ···················· | ···················· | ···················· | ···················· | ···················· |
| E | 1566 hCoV-19/Japan/PG-0469/2020\|EPI_ISL_480129\|2020-03- | ···················· | ···················· | ···················· | ···················· | ···················· | ···················· |
| E | 1567 hCoV-19/Japan/PG-0474/2020\|EPI_ISL_480134\|2020-04- | ···················· | ···················· | ···················· | ···················· | ···················· | ···················· |
| E | 1568 hCoV-19/Japan/PG-0475/2020\|EPI_ISL_480135\|2020-04- | ···················· | ···················· | ···················· | ···················· | ···················· | ···················· |
| E | 1569 hCoV-19/Japan/PG-0478/2020\|EPI_ISL_480138\|2020-04- | ···················· | ···················· | ···················· | ···················· | ···················· | ···················· |
| E | 1570 hCoV-19/Japan/PG-0480/2020\|EPI_ISL_480139\|2020-04- | ···················· | ···················· | ···················· | ···················· | ···················· | ···················· |
| E | 1571 hCoV-19/Japan/PG-0482/2020\|EPI_ISL_480141\|2020-04- | ···················· | ···················· | ···················· | ···················· | ···················· | ···················· |
| E | 1572 hCoV-19/Japan/PG-0483/2020\|EPI_ISL_480142\|2020-04- | ···················· | ···················· | ···················· | ···················· | ···················· | ···················· |
| E | 1573 hCoV-19/Japan/PG-0484/2020\|EPI_ISL_480143\|2020-04- | ···················· | ···················· | ···················· | ···················· | ···················· | ···················· |
| E | 1574 hCoV-19/Japan/PG-0485/2020\|EPI_ISL_480144\|2020-04- | ···················· | ···················· | ···················· | ···················· | ···················· | ···················· |
| E | 1575 hCoV-19/Japan/PG-0486/2020\|EPI_ISL_480145\|2020-04- | ···················· | ···················· | ···················· | ···················· | ···················· | ···················· |
| E | 1576 hCoV-19/Japan/PG-0487/2020\|EPI_ISL_480146\|2020-04- | ···················· | ···················· | ···················· | ···················· | ···················· | ···················· |
| E | 1577 hCoV-19/Japan/PG-0488/2020\|EPI_ISL_480147\|2020-04- | ···················· | ···················· | ···················· | ···················· | ···················· | ···················· |
| E | 1578 hCoV-19/Japan/PG-0489/2020\|EPI_ISL_480148\|2020-04- | ···················· | ···················· | ···················· | ···················· | ···················· | ···················· |
| E | 1579 hCoV-19/Japan/PG-0490/2020\|EPI_ISL_480149\|2020-04- | ···················· | ···················· | ···················· | ···················· | ···················· | ···················· |
| E | 1580 hCoV-19/Japan/PG-0491/2020\|EPI_ISL_480150\|2020-04- | ···················· | ···················· | ···················· | ···················· | ···················· | ···················· |
| E | 1581 hCoV-19/Japan/PG-0492/2020\|EPI_ISL_480151\|2020-04- | ···················· | ···················· | ···················· | ···················· | ···················· | ···················· |
| E | 1582 hCoV-19/Japan/PG-0493/2020\|EPI_ISL_480152\|2020-04- | ···················· | ···················· | ···················· | ···················· | ···················· | ···················· |
| E | 1583 hCoV-19/Japan/PG-0494/2020\|EPI_ISL_480153\|2020-04- | ···················· | ···················· | ···················· | ···················· | ···················· | ···················· |
| E | 1584 hCoV-19/Japan/PG-0495/2020\|EPI_ISL_480154\|2020-04- | ···················· | ···················· | ···················· | ···················· | ···················· | ···················· |
| E | 1585 hCoV-19/Japan/PG-0496/2020\|EPI_ISL_480155\|2020-04- | ···················· | ···················· | ···················· | ···················· | ···················· | ···················· |
| E | 1586 hCoV-19/Japan/PG-0497/2020\|EPI_ISL_480156\|2020-04- | ···················· | ···················· | ···················· | ···················· | ···················· | ···················· |
| E | 1587 hCoV-19/Japan/PG-0498/2020\|EPI_ISL_480157\|2020-04- | ···················· | ···················· | ···················· | ···················· | ···················· | ···················· |
| E | 1588 hCoV-19/Japan/PG-0499/2020\|EPI_ISL_480158\|2020-04- | ···················· | ···················· | ···················· | ···················· | ···················· | ···················· |
| E | 1589 hCoV-19/Japan/PG-0500/2020\|EPI_ISL_480159\|2020-04- | ···················· | ···················· | ···················· | ···················· | ···················· | ···················· |
| E | 1590 hCoV-19/Japan/PG-0501/2020\|EPI_ISL_480160\|2020-04- | ···················· | ···················· | ···················· | ···················· | ···················· | ···················· |
| E | 1591 hCoV-19/Japan/PG-0502/2020\|EPI_ISL_480161\|2020-04- | ···················· | ···················· | ···················· | ···················· | ···················· | ···················· |
| E | 1592 hCoV-19/Japan/PG-0503/2020\|EPI_ISL_480162\|2020-04- | ···················· | ···················· | ···················· | ···················· | ···················· | ···················· |
| E | 1593 hCoV-19/Japan/PG-0504/2020\|EPI_ISL_480163\|2020-04- | ···················· | ···················· | ···················· | ···················· | ···················· | ···················· |
| E | 1594 hCoV-19/Japan/PG-0505/2020\|EPI_ISL_480164\|2020-04- | ···················· | ···················· | ···················· | ···················· | ···················· | ···················· |
| E | 1595 hCoV-19/Japan/PG-0506/2020\|EPI_ISL_480165\|2020-04- | ···················· | ···················· | ···················· | ···················· | ···················· | ···················· |
| E | 1596 hCoV-19/Japan/PG-0507/2020\|EPI_ISL_480166\|2020-04- | ···················· | ···················· | ···················· | ···················· | ···················· | ···················· |
| E | 1597 hCoV-19/Japan/PG-0508/2020\|EPI_ISL_480167\|2020-04- | ···················· | ···················· | ···················· | ···················· | ···················· | ···················· |
| E | 1598 hCoV-19/Japan/PG-0509/2020\|EPI_ISL_480168\|2020-04- | ···················· | ···················· | ···················· | ···················· | ···················· | ···················· |
| E | 1599 hCoV-19/Japan/PG-0511/2020\|EPI_ISL_480169\|2020-04- | ···················· | ··········A········· | ···················· | ···················· | ···················· | ···················· |
| E | 1600 hCoV-19/Japan/PG-0512/2020\|EPI_ISL_480170\|2020-04- | ···················· | ···················· | ···················· | ···················· | ···················· | ···················· |
| E | 1601 hCoV-19/Japan/PG-0513/2020\|EPI_ISL_480171\|2020-04- | ···················· | ···················· | ···················· | ···················· | ···················· | ···················· |
| E | 1602 hCoV-19/Japan/PG-0514/2020\|EPI_ISL_480172\|2020-04- | ···················· | ···················· | ···················· | ···················· | ···················· | ···················· |
| E | 1603 hCoV-19/Japan/PG-0515/2020\|EPI_ISL_480173\|2020-04- | ···················· | ···················· | ···················· | ···················· | ···················· | ···················· |
| E | 1604 hCoV-19/Georgia/Tb-5727/2020\|EPI_ISL_481483\|2020-0 | ···················· | ···················· | ···················· | ···················· | ···················· | ···················· |
| E | 1605 hCoV-19/Finland/20A49S7/2020\|EPI_ISL_481600\|2020-0 | ···················· | ···················· | ···················· | ···················· | ···················· | ···················· |
| E | 1606 hCoV-19/Finland/20A51S7/2020\|EPI_ISL_481601\|2020-0 | ···················· | ···················· | ···················· | ···················· | ···················· | ···················· |
| E | 1607 hCoV-19/Finland/20A57S7/2020\|EPI_ISL_481602\|2020-0 | ···················· | ···················· | ···················· | ···················· | ···················· | ···················· |
| E | 1608 hCoV-19/Finland/20A69S7/2020\|EPI_ISL_481603\|2020-0 | ···················· | ···················· | ···················· | ···················· | ···················· | ···················· |
| E | 1609 hCoV-19/Finland/20A76S7/2020\|EPI_ISL_481604\|2020-0 | ···················· | ···················· | ···················· | ···················· | ···················· | ···················· |
| E | 1610 hCoV-19/Finland/21A12S11/2020\|EPI_ISL_481605\|2020- | ···················· | ···················· | ···················· | ···················· | ···················· | ···················· |
| E | 1611 hCoV-19/Finland/21A16S11/2020\|EPI_ISL_481606\|2020- | ···················· | ···················· | ···················· | ···················· | ···················· | ···················· |
| E | 1612 hCoV-19/Finland/21A19S11/2020\|EPI_ISL_481607\|2020- | ···················· | ···················· | ···················· | ···················· | ···················· | ···················· |
| E | 1613 hCoV-19/Finland/21A20S7/2020\|EPI_ISL_481608\|2020-0 | ···················· | ···················· | ···················· | ···················· | ···················· | ···················· |
| E | 1614 hCoV-19/Finland/21A27S7/2020\|EPI_ISL_481609\|2020-0 | ···················· | ···················· | ···················· | ···················· | ···················· | ···················· |
| E | 1615 hCoV-19/Finland/21A39S11/2020\|EPI_ISL_481610\|2020- | ···················· | ···················· | ···················· | ···················· | ···················· | ···················· |
| E | 1616 hCoV-19/Finland/21A41S7/2020\|EPI_ISL_481611\|2020-0 | ···················· | ···················· | ···················· | ···················· | ···················· | ···················· |
| E | 1617 hCoV-19/Finland/21A4S11/2020\|EPI_ISL_481612\|2020-0 | ···················· | ···················· | ···················· | ···················· | ···················· | ···················· |
| E | 1618 hCoV-19/Finland/21A50S11/2020\|EPI_ISL_481613\|2020- | ···················· | ···················· | ···················· | ···················· | ···················· | ···················· |
| E | 1619 hCoV-19/Finland/22A33S5/2020\|EPI_ISL_481614\|2020-0 | ···················· | ···················· | ···················· | ···················· | ···················· | ···················· |
| E | 1620 hCoV-19/Finland/22A44S5/2020\|EPI_ISL_481615\|2020-0 | ···················· | ···················· | ···················· | ···················· | ···················· | ···················· |
| E | 1621 hCoV-19/Finland/22A45S8/2020\|EPI_ISL_481616\|2020-0 | ···················· | ···················· | ···················· | ···················· | ···················· | ···················· |
| E | 1622 hCoV-19/Finland/22A98S8/2020\|EPI_ISL_481617\|2020-0 | ···················· | ···················· | ···················· | ···················· | ···················· | ···················· |
| E | 1623 hCoV-19/Finland/26A24S5/2020\|EPI_ISL_481618\|2020-0 | ···················· | ···················· | ···················· | ···················· | ···················· | ···················· |
| E | 1624 hCoV-19/Finland/26A44S5/2020\|EPI_ISL_481619\|2020-0 | ···················· | ···················· | ···················· | ···················· | ···················· | ···················· |
| E | 1625 hCoV-19/South Africa/KRISP-0191/2020\|EPI_ISL_48271 | ···················· | ···················· | ···················· | ···················· | ···················· | ···················· |
| E | 1626 hCoV-19/South Africa/KRISP-0192/2020\|EPI_ISL_48271 | ···················· | ···················· | ···················· | ···················· | ···················· | ···················· |
| E | 1627 hCoV-19/South Africa/KRISP-0196/2020\|EPI_ISL_48271 | ···················· | ···················· | ···················· | ···················· | ···················· | ···················· |
| E | 1628 hCoV-19/South Africa/KRISP-0201/2020\|EPI_ISL_48271 | ···················· | ···················· | ···················· | ···················· | ···················· | ···················· |
| E | 1629 hCoV-19/South Africa/KRISP-0202/2020\|EPI_ISL_48271 | ···················· | ···················· | ···················· | ···················· | ···················· | ···················· |
| E | 1630 hCoV-19/South Africa/KRISP-0205/2020\|EPI_ISL_48271 | ···················· | ···················· | ···················· | ···················· | ···················· | ···················· |
| E | 1631 hCoV-19/South Africa/KRISP-0206/2020\|EPI_ISL_48271 | ···················· | ···················· | ···················· | ···················· | ···················· | ···················· |
| E | 1632 hCoV-19/South Africa/KRISP-0211/2020\|EPI_ISL_48272 | ···················· | ···················· | ···················· | ···················· | ···················· | ···················· |
| E | 1633 hCoV-19/South Africa/KRISP-0212/2020\|EPI_ISL_48272 | ···················· | ···················· | ···················· | ···················· | ···················· | ···················· |
| E | 1634 hCoV-19/South Africa/KRISP-0213/2020\|EPI_ISL_48272 | ···················· | ···················· | ···················· | ···················· | ···················· | ···················· |
| E | 1635 hCoV-19/South Africa/KRISP-0214/2020\|EPI_ISL_48272 | ···················· | ···················· | ···················· | ···················· | ···················· | ···················· |
| E | 1636 hCoV-19/South Africa/KRISP-0245/2020\|EPI_ISL_48272 | ···················· | ···················· | ···················· | ···················· | ···················· | ···················· |

| | | | | | | | | |
|---|---|---|---|---|---|---|---|---|
| E | 1637 | hCoV-19/Serbia/UE92/2020\|EPI_ISL_482744\|2020-04-03 | ···················· | ···················· | ···················· | ···················· | ···················· | ···················· |
| E | 1638 | hCoV-19/Netherlands/Leiden-0008/2020\|EPI_ISL_48274 | ···················· | ···················· | ···················· | ···················· | ···················· | ···················· |
| E | 1639 | hCoV-19/Netherlands/Leiden-0001/2020\|EPI_ISL_48274 | ···················· | ···················· | ···················· | ···················· | ···················· | ···················· |
| E | 1640 | hCoV-19/Egypt/MASRI-005/2020\|EPI_ISL_482759\|2020-0 | ···················· | ···················· | ···················· | ···················· | ···················· | ···················· |
| E | 1644 | hCoV-19/Egypt/MASRI-012/2020\|EPI_ISL_482763\|2020-0 | ···················· | ···················· | ···················· | ···················· | ···················· | ···················· |
| E | 1645 | hCoV-19/Egypt/MASRI-013/2020\|EPI_ISL_482764\|2020-0 | ···················· | ···················· | ···················· | ···················· | ···················· | ···················· |
| E | 1647 | hCoV-19/Egypt/MASRI-015/2020\|EPI_ISL_482766\|2020-0 | ···················· | ·A·················· | ···················· | ···················· | ···················· | ···A················ |
| E | 1649 | hCoV-19/Egypt/MASRI-022/2020\|EPI_ISL_482768\|2020-0 | ···················· | ···················· | ···················· | ···················· | ···················· | ···················· |
| E | 1650 | hCoV-19/Egypt/MASRI-023/2020\|EPI_ISL_482769\|2020-0 | ···················· | ···················· | ···················· | ···················· | ···················· | ···················· |
| E | 1651 | hCoV-19/Egypt/MASRI-024/2020\|EPI_ISL_482770\|2020-0 | ···················· | ······A············· | ···················· | ···················· | ···················· | ···················· |
| E | 1653 | hCoV-19/Egypt/MASRI-026/2020\|EPI_ISL_482772\|2020-0 | ···················· | ···················· | ···················· | ···················· | ···················· | ···················· |
| E | 1654 | hCoV-19/Egypt/MASRI-027/2020\|EPI_ISL_482773\|2020-0 | ···················· | ···················· | ···················· | ···················· | ···················· | ···················· |
| E | 1655 | hCoV-19/Egypt/MASRI-029/2020\|EPI_ISL_482774\|2020-0 | ···················· | ······A············· | ···················· | ···················· | ···················· | ···················· |
| E | 1656 | hCoV-19/USA/MN-MDH-1225/2020\|EPI_ISL_482946\|2020-0 | ···················· | ···················· | ···················· | ···················· | ···················· | ···················· |
| E | 1657 | hCoV-19/USA/MN-MDH-1226/2020\|EPI_ISL_482947\|2020-0 | ···················· | ···················· | ···················· | ···················· | ···················· | ···················· |
| E | 1658 | hCoV-19/USA/MN-MDH-1227/2020\|EPI_ISL_482948\|2020-0 | ···················· | ···················· | ···················· | ···················· | ···················· | ···················· |
| E | 1659 | hCoV-19/USA/MN-MDH-1228/2020\|EPI_ISL_482949\|2020-0 | ···················· | ···················· | ···················· | ···················· | ···················· | ···················· |
| E | 1660 | hCoV-19/USA/LA-GBCL-26544587/2020\|EPI_ISL_485156\|2 | ···················· | ···················· | ···················· | ···················· | ···················· | ···················· |
| E | 1661 | hCoV-19/USA/LA-GBCL-26556560/2020\|EPI_ISL_485190\|2 | ···················· | ···················· | ···················· | ···················· | ···················· | ···················· |
| E | 1662 | hCoV-19/USA/LA-GBCL-26556561/2020\|EPI_ISL_485191\|2 | ···················· | ···················· | ···················· | ···················· | ···················· | ···················· |
| E | 1663 | hCoV-19/USA/LA-GBCL-26556562/2020\|EPI_ISL_485192\|2 | ···················· | ···················· | ···················· | ···················· | ···················· | ···················· |
| E | 1664 | hCoV-19/USA/LA-GBCL-26556563/2020\|EPI_ISL_485193\|2 | ···················· | ···················· | ···················· | ···················· | ···················· | ···················· |
| E | 1665 | hCoV-19/USA/LA-GBCL-26556564/2020\|EPI_ISL_485194\|2 | ···················· | ···················· | ···················· | ···················· | ···················· | ···················· |
| E | 1666 | hCoV-19/USA/LA-GBCL-26556565/2020\|EPI_ISL_485195\|2 | ···················· | ···················· | ···················· | ···················· | ···················· | ···················· |
| E | 1667 | hCoV-19/USA/LA-GBCL-26556567/2020\|EPI_ISL_485197\|2 | ···················· | ···················· | ···················· | ···················· | ···················· | ···················· |
| E | 1668 | hCoV-19/USA/LA-GBCL-26556569/2020\|EPI_ISL_485198\|2 | ···················· | ···················· | ···················· | ···················· | ···················· | ···················· |
| E | 1669 | hCoV-19/USA/LA-GBCL-26556570/2020\|EPI_ISL_485199\|2 | ···················· | ···················· | ···················· | ···················· | ···················· | ···················· |
| E | 1672 | hCoV-19/Switzerland/ZH-ETHZ-110057/2020\|EPI_ISL_48 | ···················· | ···················· | ···················· | ···················· | ···················· | ···················· |
| E | 1673 | hCoV-19/Australia/SAP119/2020\|EPI_ISL_492109\|2020- | ···················· | ···················· | ···················· | ···················· | ···················· | ···················· |
| E | 1674 | hCoV-19/Australia/SAP121/2020\|EPI_ISL_492110\|2020- | ···················· | ···················· | ···················· | ···················· | ···················· | ···················· |
| E | 1675 | hCoV-19/Australia/SAP123/2020\|EPI_ISL_492111\|2020- | ···················· | ···················· | ···················· | ···················· | ···················· | ···················· |
| E | 1676 | hCoV-19/Australia/SAP151/2020\|EPI_ISL_492113\|2020- | ···················· | ···················· | ···················· | ···················· | ···················· | ···················· |
| E | 1677 | hCoV-19/Australia/SAP159/2020\|EPI_ISL_492116\|2020- | ···················· | ···················· | ···················· | ···················· | ···················· | ···················· |
| E | 1678 | hCoV-19/Australia/SAP161/2020\|EPI_ISL_492117\|2020- | ···················· | ···················· | ···················· | ···················· | ···················· | ···················· |
| E | 1679 | hCoV-19/Australia/SAP197/2020\|EPI_ISL_492130\|2020- | ···················· | ···················· | ···················· | ···················· | ···················· | ···················· |
| E | 1680 | hCoV-19/Australia/SAP198/2020\|EPI_ISL_492131\|2020- | ···················· | ···················· | ···················· | ···················· | ···················· | ···················· |
| E | 1681 | hCoV-19/Australia/SAP368/2020\|EPI_ISL_492133\|2020- | ···················· | ···················· | ···················· | ···················· | ···················· | ···················· |
| E | 1682 | hCoV-19/Australia/SAP369/2020\|EPI_ISL_492134\|2020- | ···················· | ···················· | ···················· | ···················· | ···················· | ···················· |
| E | 1683 | hCoV-19/Australia/SAP370/2020\|EPI_ISL_492135\|2020- | ···················· | ···················· | ···················· | ···················· | ···················· | ···················· |
| E | 1684 | hCoV-19/Australia/SAP371/2020\|EPI_ISL_492136\|2020- | ········T··········· | ···················· | ···················· | ···················· | ···················· | ···················· |
| E | 1685 | hCoV-19/Australia/SAP372/2020\|EPI_ISL_492137\|2020- | ···················· | ···················· | ···················· | ···················· | ···················· | ···················· |
| E | 1686 | hCoV-19/Australia/SAP373/2020\|EPI_ISL_492138\|2020- | ···················· | ···················· | ···················· | ···················· | ···················· | ···················· |
| E | 1687 | hCoV-19/Australia/SAP375/2020\|EPI_ISL_492140\|2020- | ···················· | ···················· | ···················· | ···················· | ···················· | ···················· |
| E | 1688 | hCoV-19/Australia/SAP376/2020\|EPI_ISL_492141\|2020- | ···················· | ···················· | ···················· | ···················· | ···················· | ···················· |
| E | 1689 | hCoV-19/Australia/SAP377/2020\|EPI_ISL_492142\|2020- | ···················· | ···················· | ···················· | ···················· | ···················· | ···················· |
| E | 1690 | hCoV-19/Australia/SAP378/2020\|EPI_ISL_492143\|2020- | ···················· | ···················· | ···················· | ···················· | ···················· | ···················· |
| E | 1691 | hCoV-19/Australia/SAP379/2020\|EPI_ISL_492144\|2020- | ···················· | ···················· | ···················· | ···················· | ···················· | ···················· |
| E | 1692 | hCoV-19/Australia/SAP380/2020\|EPI_ISL_492145\|2020- | ···················· | ···················· | ···················· | ···················· | ···················· | ···················· |
| E | 1693 | hCoV-19/Australia/SAP381/2020\|EPI_ISL_492146\|2020- | ···················· | ···················· | ···················· | ···················· | ···················· | ···················· |
| E | 1694 | hCoV-19/Australia/SAP382/2020\|EPI_ISL_492147\|2020- | ···················· | ···················· | ···················· | ···················· | ···················· | ···················· |
| E | 1695 | hCoV-19/Australia/SAP383/2020\|EPI_ISL_492148\|2020- | ···················· | ···················· | ···················· | ···················· | ···················· | ···················· |
| E | 1696 | hCoV-19/Australia/SAP384/2020\|EPI_ISL_492149\|2020- | ···················· | ···················· | ···················· | ···················· | ···················· | ···················· |
| E | 1697 | hCoV-19/Australia/SAP385/2020\|EPI_ISL_492150\|2020- | ···················· | ···················· | ···················· | ···················· | ···················· | ···················· |
| E | 1698 | hCoV-19/Australia/SAP386/2020\|EPI_ISL_492151\|2020- | ···················· | ···················· | ···················· | ···················· | ···················· | ···················· |
| E | 1699 | hCoV-19/Australia/SAP387/2020\|EPI_ISL_492152\|2020- | ···················· | ···················· | ···················· | ···················· | ···················· | ···················· |
| E | 1701 | hCoV-19/Singapore/648/2020\|EPI_ISL_493408\|2020-04- | ···················· | ···················· | ···················· | ···················· | ···················· | ···················· |
| E | 1702 | hCoV-19/Singapore/649/2020\|EPI_ISL_493409\|2020-05- | ···················· | ···················· | ···················· | ···················· | ···················· | ···················· |
| E | 1703 | hCoV-19/Singapore/650/2020\|EPI_ISL_493410\|2020-04- | ···················· | ···················· | ···················· | ···················· | ···················· | ···················· |
| E | 1704 | hCoV-19/Singapore/651/2020\|EPI_ISL_493411\|2020-05- | ···················· | ···················· | ···················· | ···················· | ···················· | ···················· |
| E | 1705 | hCoV-19/Singapore/652/2020\|EPI_ISL_493412\|2020-04- | ···················· | ···················· | ···················· | ···················· | ···················· | ···················· |
| E | 1706 | hCoV-19/Singapore/653/2020\|EPI_ISL_493413\|2020-04- | ···················· | ···················· | ···················· | ···················· | ···················· | ···················· |
| E | 1707 | hCoV-19/Singapore/654/2020\|EPI_ISL_493414\|2020-02- | ···················· | ···················· | ···················· | ···················· | ···················· | ···················· |
| E | 1708 | hCoV-19/Singapore/655/2020\|EPI_ISL_493415\|2020-03- | ···················· | ···················· | ···················· | ···················· | ···················· | ···················· |
| E | 1709 | hCoV-19/Singapore/656/2020\|EPI_ISL_493416\|2020-04- | ···················· | ···················· | ···················· | ···················· | ···················· | ···················· |
| E | 1710 | hCoV-19/Singapore/657/2020\|EPI_ISL_493417\|2020-03- | ···················· | ···················· | ···················· | ···················· | ···················· | ···················· |
| E | 1711 | hCoV-19/Singapore/658/2020\|EPI_ISL_493418\|2020-04- | ···················· | ···················· | ···················· | ···················· | ···················· | ···················· |
| E | 1712 | hCoV-19/Singapore/659/2020\|EPI_ISL_493419\|2020-03- | ···················· | ···················· | ···················· | ···················· | ···················· | ···················· |
| E | 1713 | hCoV-19/Singapore/660/2020\|EPI_ISL_493420\|2020-04- | ···················· | ···················· | ···················· | ···················· | ···················· | ···················· |
| E | 1714 | hCoV-19/Singapore/661/2020\|EPI_ISL_493421\|2020-02- | ···················· | ···················· | ···················· | ···················· | ···················· | ···················· |
| E | 1715 | hCoV-19/Singapore/662/2020\|EPI_ISL_493422\|2020-04- | ···················· | ···················· | ···················· | ···················· | ···················· | ···················· |
| E | 1716 | hCoV-19/Singapore/663/2020\|EPI_ISL_493423\|2020-04- | ···················· | ···················· | ···················· | ···················· | ···················· | ···················· |
| E | 1717 | hCoV-19/Singapore/664/2020\|EPI_ISL_493424\|2020-04- | ···················· | ···················· | ···················· | ···················· | ···················· | ···················· |
| E | 1718 | hCoV-19/Singapore/665/2020\|EPI_ISL_493425\|2020-03- | ···················· | ···················· | ···················· | ···················· | ···················· | ···················· |
| E | 1719 | hCoV-19/USA/SC-USC-USC00431/2020\|EPI_ISL_493426\|20 | ···················· | ··············C····· | ···················· | ···················· | ···················· | ···················· |
| E | 1720 | hCoV-19/USA/SC-USC-PRG443/2020\|EPI_ISL_493427\|2020 | ···················· | ···················· | ···················· | ···················· | ···················· | ···················· |
| E | 1721 | hCoV-19/England/BIRM-6000E/2020\|EPI_ISL_493429\|202 | ···················· | ···················· | ···················· | ···················· | ···················· | ···················· |
| E | 1722 | hCoV-19/England/BIRM-6004A/2020\|EPI_ISL_493432\|202 | ···················· | ···················· | ···················· | ···················· | ···················· | ···················· |
| E | 1723 | hCoV-19/England/BIRM-600B3/2020\|EPI_ISL_493437\|202 | ···················· | ···················· | ···················· | ···················· | ···················· | ···················· |
| E | 1724 | hCoV-19/England/BIRM-6011A/2020\|EPI_ISL_493439\|202 | ···················· | ···················· | ···················· | ···················· | ···················· | ···················· |
| E | 1725 | hCoV-19/England/BIRM-6161A/2020\|EPI_ISL_493444\|202 | ···················· | ···················· | ···················· | ···················· | ···················· | ···················· |

| | | | |
|---|---|---|---|
| E | 1726 | hCoV-19/England/BIRM-61629/2020\|EPI_ISL_493445\|202 | .................... .................... ..................... .................... .................... .................... |
| E | 1727 | hCoV-19/England/BIRM-61656/2020\|EPI_ISL_493447\|202 | .................... .................... ..................... .................... .................... .................... |
| E | 1728 | hCoV-19/England/BIRM-61665/2020\|EPI_ISL_493448\|202 | .................... .................... ..................... .................... .................... .................... |
| E | 1729 | hCoV-19/England/BIRM-61674/2020\|EPI_ISL_493449\|202 | .................... .................... ..................... .................... .................... .................... |
| E | 1730 | hCoV-19/England/NORT-285C67/2020\|EPI_ISL_493460\|20 | .................... .................... ..................... .................... .................... .................... |
| E | 1731 | hCoV-19/England/NORT-285CD0/2020\|EPI_ISL_493461\|20 | .................... .................... ..................... .................... .................... .................... |
| E | 1732 | hCoV-19/England/NORT-285D64/2020\|EPI_ISL_493463\|20 | .................... .................... ..................... .................... .................... .................... |
| E | 1733 | hCoV-19/England/NORT-285DBF/2020\|EPI_ISL_493464\|20 | .................... .................... ..................... .................... .................... .................... |
| E | 1734 | hCoV-19/England/NORT-285E61/2020\|EPI_ISL_493465\|20 | .................... .................... ..................... .................... .................... .................... |
| E | 1735 | hCoV-19/England/NORT-285F04/2020\|EPI_ISL_493466\|20 | .................... .................... ..................... .................... .................... .................... |
| E | 1736 | hCoV-19/England/NORT-298821/2020\|EPI_ISL_493468\|20 | .................... .................... ..................... .................... .................... .................... |
| E | 1737 | hCoV-19/England/NORT-298830/2020\|EPI_ISL_493469\|20 | .................... .................... ..................... .................... .................... .................... |
| E | 1738 | hCoV-19/England/NORT-29885E/2020\|EPI_ISL_493470\|20 | .................... .................... ..................... .................... .................... .................... |
| E | 1739 | hCoV-19/England/NORT-298AD0/2020\|EPI_ISL_493472\|20 | .................... .................... ..................... .................... .................... .................... |
| E | 1740 | hCoV-19/England/NORT-298DAA/2020\|EPI_ISL_493476\|20 | .................... .................... ..................... .................... .................... .................... |
| E | 1741 | hCoV-19/England/NORT-298E01/2020\|EPI_ISL_493477\|20 | .................... .................... ..................... .................... .................... .................... |
| E | 1742 | hCoV-19/England/NORT-298ED4/2020\|EPI_ISL_493478\|20 | .................... .................... ..................... .................... .................... .................... |
| E | 1743 | hCoV-19/England/NORT-299019/2020\|EPI_ISL_493480\|20 | .................... .................... ..................... .................... .................... .................... |
| E | 1744 | hCoV-19/England/NORT-299170/2020\|EPI_ISL_493481\|20 | .................... .................... ..................... .................... .................... .................... |
| E | 1745 | hCoV-19/England/NORT-2991AD/2020\|EPI_ISL_493482\|20 | .................... .................... ..................... .................... .................... .................... |
| E | 1746 | hCoV-19/USA/CA-ALSR-1630/2020\|EPI_ISL_494700\|2020- | .................... .................... ..................... .................... .................... .................... |
| E | 1747 | hCoV-19/USA/CA-ALSR-1631/2020\|EPI_ISL_494701\|2020- | .................... .A.................. ..................... .................... .................... .................... |
| E | 1748 | hCoV-19/USA/CA-ALSR-1632/2020\|EPI_ISL_494702\|2020- | .................... .A.................. ..................... .................... .................... .................... |
| E | 1749 | hCoV-19/USA/CA-ALSR-1633/2020\|EPI_ISL_494703\|2020- | .................... .A.................. ..................... .................... .................... .................... |
| E | 1750 | hCoV-19/USA/CA-ALSR-1634/2020\|EPI_ISL_494704\|2020- | .................... .................... ..................... .................... .................... .................... |
| E | 1751 | hCoV-19/USA/CA-ALSR-1636/2020\|EPI_ISL_494705\|2020- | .................... .................... ..................... .................... .................... .................... |
| E | 1752 | hCoV-19/USA/CA-ALSR-1638/2020\|EPI_ISL_494706\|2020- | .................... .................... ..................... .................... .................... .................... |
| E | 1753 | hCoV-19/USA/CA-ALSR-1640/2020\|EPI_ISL_494707\|2020- | .................... .................... ..................... .................... .................... .................... |
| E | 1754 | hCoV-19/USA/CA-ALSR-1641/2020\|EPI_ISL_494708\|2020- | .................... .................... ..................... .................... .................... .................... |
| E | 1755 | hCoV-19/USA/CA-ALSR-1643/2020\|EPI_ISL_494709\|2020- | .................... .................... ..................... .................... .................... .................... |
| E | 1756 | hCoV-19/USA/CA-ALSR-1645/2020\|EPI_ISL_494710\|2020- | .................... .................... ..................... .................... .................... .................... |
| E | 1757 | hCoV-19/USA/CA-ALSR-1647/2020\|EPI_ISL_494712\|2020- | .................... .................... ..................... .................... .................... .................... |
| E | 1758 | hCoV-19/USA/CA-ALSR-1649/2020\|EPI_ISL_494714\|2020- | .................... .A.................. ..................... .................... .................... .................... |
| E | 1759 | hCoV-19/USA/CA-ALSR-1651/2020\|EPI_ISL_494716\|2020- | .................... .................... ..................... .................... .................... .................... |
| E | 1760 | hCoV-19/USA/CA-ALSR-1653/2020\|EPI_ISL_494717\|2020- | .................... .................... ..................... .................... .................... .................... |
| E | 1761 | hCoV-19/USA/CA-ALSR-1654/2020\|EPI_ISL_494718\|2020- | .................... .................... ..................... .................... .................... .................... |
| E | 1762 | hCoV-19/USA/CA-ALSR-1657/2020\|EPI_ISL_494719\|2020- | .................... .................... ..................... .................... .................... .................... |
| E | 1763 | hCoV-19/USA/CA-ALSR-1658/2020\|EPI_ISL_494720\|2020- | .................... .................... ..................... .................... .................... .................... |
| E | 1764 | hCoV-19/USA/CA-ALSR-1660/2020\|EPI_ISL_494721\|2020- | .................... .................... ..................... .................... .................... .................... |
| E | 1765 | hCoV-19/USA/CA-ALSR-1661/2020\|EPI_ISL_494722\|2020- | .................... .................... ..................... .................... .................... .................... |
| E | 1766 | hCoV-19/USA/CA-ALSR-1662/2020\|EPI_ISL_494723\|2020- | .................... .................... ..................... .................... .................... .................... |
| E | 1767 | hCoV-19/USA/CA-ALSR-1663/2020\|EPI_ISL_494724\|2020- | .................... .................... ..................... .................... .................... .................... |
| E | 1768 | hCoV-19/USA/CA-ALSR-1664/2020\|EPI_ISL_494725\|2020- | .................... .................... ..................... .................... .................... .................... |
| E | 1769 | hCoV-19/USA/CA-ALSR-1665/2020\|EPI_ISL_494726\|2020- | .................... .................... ..................... .................... .................... .................... |
| E | 1770 | hCoV-19/USA/CA-ALSR-1667/2020\|EPI_ISL_494727\|2020- | .................... .................... ..................... .................... .................... .................... |
| E | 1771 | hCoV-19/USA/CA-ALSR-1668/2020\|EPI_ISL_494728\|2020- | .................... .................... ..................... .................... .................... .................... |
| E | 1772 | hCoV-19/USA/CA-ALSR-1669/2020\|EPI_ISL_494729\|2020- | .................... .................... ..................... .................... .................... .................... |
| E | 1773 | hCoV-19/USA/CA-ALSR-1670/2020\|EPI_ISL_494730\|2020- | .................... .................... ..................... .................... .................... .................... |
| E | 1774 | hCoV-19/USA/CA-ALSR-1671/2020\|EPI_ISL_494731\|2020- | .................... .................... ..................... .................... .................... .................... |
| E | 1775 | hCoV-19/USA/CA-ALSR-1672/2020\|EPI_ISL_494732\|2020- | .................... .................... ..................... .................... .................... .................... |
| E | 1776 | hCoV-19/USA/CA-ALSR-1673/2020\|EPI_ISL_494733\|2020- | .................... .................... ..................... .................... .................... .................... |
| E | 1777 | hCoV-19/USA/CA-ALSR-1674/2020\|EPI_ISL_494734\|2020- | .................... .................... ..................... .................... .................... .................... |
| E | 1778 | hCoV-19/USA/CA-ALSR-1675/2020\|EPI_ISL_494735\|2020- | .................... .................... ..................... .................... .................... .................... |
| E | 1779 | hCoV-19/USA/CA-ALSR-1676/2020\|EPI_ISL_494736\|2020- | .................... .................... ..................... .................... .................... .................... |
| E | 1780 | hCoV-19/USA/CA-ALSR-1677/2020\|EPI_ISL_494737\|2020- | .................... .................... ..................... .................... .................... .................... |
| E | 1781 | hCoV-19/USA/CA-ALSR-1681/2020\|EPI_ISL_494739\|2020- | .................... .................... ..................... .................... .................... .................... |
| E | 1782 | hCoV-19/USA/CA-ALSR-1682/2020\|EPI_ISL_494740\|2020- | .................... .................... ..................... .................... .................... .................... |
| E | 1783 | hCoV-19/USA/CA-ALSR-1683/2020\|EPI_ISL_494741\|2020- | .................... .................... ..................... .................... .................... .................... |
| E | 1784 | hCoV-19/USA/CA-ALSR-1684/2020\|EPI_ISL_494742\|2020- | .................... .................... ..................... .................... .................... .................... |
| E | 1785 | hCoV-19/USA/CA-ALSR-1685/2020\|EPI_ISL_494743\|2020- | .................... .................... ..................... .................... .................... .................... |
| E | 1786 | hCoV-19/USA/CA-ALSR-1687/2020\|EPI_ISL_494744\|2020- | .................... .................... ..................... .................... .................... .................... |
| E | 1787 | hCoV-19/USA/CA-ALSR-1688/2020\|EPI_ISL_494745\|2020- | .................... .A.................. ..................... .................... .................... .................... |
| E | 1788 | hCoV-19/USA/CA-ALSR-1689/2020\|EPI_ISL_494746\|2020- | .................... .A.................. ..................... .................... .................... .................... |
| E | 1789 | hCoV-19/Italy/CAM-INTPas-5/2020\|EPI_ISL_494757\|202 | .................... .................... ..................... .................... .................... .................... |
| E | 1790 | hCoV-19/Italy/CAM-INTPas-6/2020\|EPI_ISL_494759\|202 | .................... .................... ..................... .................... .................... .................... |
| E | 1791 | hCoV-19/Italy/CAM-INTPas-7/2020\|EPI_ISL_494761\|202 | .................... .................... ..................... .................... .................... .................... |
| E | 1792 | hCoV-19/Italy/CAM-INTPas-10/2020\|EPI_ISL_494771\|20 | .................... .................... ..................... .................... .................... .................... |
| E | 1793 | hCoV-19/Turkey/IST-GLAB-152/2020\|EPI_ISL_480251\|20 | .................... .................... ..................... .................... .................... .................... |
| E | 1794 | hCoV-19/Turkey/IST-GLAB-153/2020\|EPI_ISL_480252\|20 | .................... .................... ..................... .................... .................... .................... |
| E | 1795 | hCoV-19/Turkey/IST-GLAB-155/2020\|EPI_ISL_480253\|20 | .................... .................... ..................... .................... .................... .................... |
| E | 1796 | hCoV-19/Turkey/IST-GLAB-156/2020\|EPI_ISL_480254\|20 | .................... .................... ..................... .................... .................... .................... |
| E | 1797 | hCoV-19/Turkey/IST-GLAB-157/2020\|EPI_ISL_480255\|20 | .................... .................... ..................... .................... .................... .................... |
| E | 1798 | hCoV-19/Turkey/IST-GLAB-158/2020\|EPI_ISL_480256\|20 | .................... .................... ..................... .................... .................... .................... |
| E | 1799 | hCoV-19/Turkey/IST-GLAB-162/2020\|EPI_ISL_480257\|20 | .................... .................... ..................... .................... .................... .................... |
| E | 1800 | hCoV-19/Turkey/IST-GLAB-163/2020\|EPI_ISL_480258\|20 | .................... .................... ..................... .................... .................... .................... |
| E | 1801 | hCoV-19/Turkey/IST-GLAB-164/2020\|EPI_ISL_480259\|20 | .................... .................... ..................... .................... .................... .................... |
| E | 1802 | hCoV-19/Turkey/IST-GLAB-167/2020\|EPI_ISL_480260\|20 | .................... .................... ..................... .................... .................... .................... |
| E | 1803 | hCoV-19/Turkey/IST-GLAB-168/2020\|EPI_ISL_480261\|20 | .................... .................... ..................... .................... .................... .................... |
| E | 1804 | hCoV-19/Bulgaria/06/2020\|EPI_ISL_480297\|2020-03-21 | .................... .................... ..................... .................... .................... .................... |
| E | 1805 | hCoV-19/Bulgaria/15/2020\|EPI_ISL_480298\|2020-05-13 | .................... .................... ..................... .................... .................... .................... |

| | | | | | | | |
|---|---|---|---|---|---|---|---|
| E | 1806 hCoV-19/Bulgaria/16/2020\|EPI_ISL_480299\|2020-04-26 | .................... | .................... | ..................... | .................... | .................... | .................... |
| E | 1807 hCoV-19/Finland/17A66S12/2020\|EPI_ISL_481582\|2020- | .................... | .................... | ..................... | .................... | .................... | .................... |
| E | 1808 hCoV-19/Finland/17A69S12/2020\|EPI_ISL_481583\|2020- | .................... | .................... | ..................... | .................... | .................... | .................... |
| E | 1809 hCoV-19/Finland/17A70S12/2020\|EPI_ISL_481584\|2020- | .................... | .................... | ..................... | .................... | .................... | .................... |
| E | 1810 hCoV-19/Finland/17A73S13/2020\|EPI_ISL_481585\|2020- | .................... | .................... | ..................... | .................... | .................... | .................... |
| E | 1811 hCoV-19/Finland/17A76S12/2020\|EPI_ISL_481586\|2020- | .................... | .................... | ..................... | .................... | .................... | .................... |
| E | 1812 hCoV-19/Finland/17A7S13/2020\|EPI_ISL_481587\|2020-0 | .................... | .................... | ..................... | .................... | .................... | .................... |
| E | 1813 hCoV-19/Finland/17A82S13/2020\|EPI_ISL_481588\|2020- | .................... | .................... | ..................... | .................... | .................... | .................... |
| E | 1814 hCoV-19/Finland/17A85S10/2020\|EPI_ISL_481589\|2020- | .................... | .................... | ..................... | .................... | .................... | ...............A.... |
| E | 1815 hCoV-19/Finland/17A87S10/2020\|EPI_ISL_481590\|2020- | .................... | .................... | ..................... | .................... | .................... | .................... |
| E | 1816 hCoV-19/Finland/17A91S12/2020\|EPI_ISL_481591\|2020- | .................... | .................... | ..................... | .................... | .................... | .................... |
| E | 1817 hCoV-19/Finland/17A91S13/2020\|EPI_ISL_481592\|2020- | .................... | .................... | ..................... | .................... | .................... | .................... |
| E | 1818 hCoV-19/USA/LA-GBCL-26544503/2020\|EPI_ISL_485101\|2 | .................... | .................... | ..................... | .................... | .................... | .................... |
| E | 1819 hCoV-19/USA/LA-GBCL-26544505/2020\|EPI_ISL_485103\|2 | .................... | .................... | ..................... | .................... | .................... | .................... |
| E | 1820 hCoV-19/USA/LA-GBCL-26544513/2020\|EPI_ISL_485105\|2 | .................... | .................... | ..................... | .................... | .................... | .................... |
| E | 1821 hCoV-19/USA/LA-GBCL-26544521/2020\|EPI_ISL_485108\|2 | .................... | .................... | ..................... | .................... | .................... | .................... |
| E | 1822 hCoV-19/USA/LA-GBCL-26544523/2020\|EPI_ISL_485109\|2 | .................... | .................... | ..................... | .................... | .................... | .................... |
| E | 1823 hCoV-19/USA/LA-GBCL-26544524/2020\|EPI_ISL_485110\|2 | .................... | .................... | ..................... | .................... | .................... | .................... |
| E | 1824 hCoV-19/USA/LA-GBCL-26544532/2020\|EPI_ISL_485115\|2 | .................... | .................... | ..................... | .................... | .................... | .................... |
| E | 1825 hCoV-19/USA/LA-GBCL-26544536/2020\|EPI_ISL_485118\|2 | .................... | .................... | ..................... | .................... | .................... | .................... |
| E | 1826 hCoV-19/USA/LA-GBCL-26544538/2020\|EPI_ISL_485119\|2 | .................... | .................... | ..................... | .................... | .................... | .................... |
| E | 1827 hCoV-19/USA/LA-GBCL-26544539/2020\|EPI_ISL_485120\|2 | .................... | .................... | ..................... | .................... | .................... | .................... |
| E | 1828 hCoV-19/USA/LA-GBCL-26544542/2020\|EPI_ISL_485123\|2 | .................... | .................... | ..................... | .................... | .................... | .................... |
| E | 1829 hCoV-19/USA/LA-GBCL-26544543/2020\|EPI_ISL_485124\|2 | .................... | .................... | ..................... | .................... | .................... | .................... |
| E | 1830 hCoV-19/USA/LA-GBCL-26544548/2020\|EPI_ISL_485127\|2 | .................... | .................... | ..................... | .................... | .................... | .................... |
| E | 1831 hCoV-19/USA/LA-GBCL-26544551/2020\|EPI_ISL_485129\|2 | .................... | .................... | ..................... | .................... | .................... | .................... |
| E | 1832 hCoV-19/USA/LA-GBCL-26544561/2020\|EPI_ISL_485136\|2 | .................... | .................... | ..................... | .................... | .................... | .................... |
| E | 1833 hCoV-19/USA/LA-GBCL-26544565/2020\|EPI_ISL_485138\|2 | .................... | .................... | ..................... | .................... | .................... | .................... |
| E | 1834 hCoV-19/USA/LA-GBCL-26544567/2020\|EPI_ISL_485140\|2 | .................... | .................... | ..................... | .................... | .................... | .................... |
| E | 1835 hCoV-19/USA/LA-GBCL-26544569/2020\|EPI_ISL_485141\|2 | .................... | .................... | ..................... | .................... | .................... | .................... |
| E | 1836 hCoV-19/USA/LA-GBCL-26544570/2020\|EPI_ISL_485142\|2 | .................... | .................... | ..................... | .................... | .................... | .................... |
| E | 1837 hCoV-19/USA/LA-GBCL-26544573/2020\|EPI_ISL_485145\|2 | .................... | .................... | ..................... | .................... | .................... | .................... |
| E | 1838 hCoV-19/USA/LA-GBCL-26544574/2020\|EPI_ISL_485146\|2 | .................... | .................... | ..................... | .................... | .................... | .................... |
| E | 1839 hCoV-19/USA/LA-GBCL-26544575/2020\|EPI_ISL_485147\|2 | .................... | .................... | ..................... | .................... | .................... | .................... |
| E | 1840 hCoV-19/USA/LA-GBCL-26544576/2020\|EPI_ISL_485148\|2 | .................... | .................... | ..................... | .................... | .................... | .................... |
| E | 1841 hCoV-19/USA/LA-GBCL-26544579/2020\|EPI_ISL_485150\|2 | .................... | .................... | ..................... | .................... | .................... | .................... |
| E | 1842 hCoV-19/USA/LA-GBCL-26556502/2020\|EPI_ISL_485157\|2 | .................... | .................... | ..................... | .................... | .................... | .................... |
| E | 1843 hCoV-19/USA/LA-GBCL-26556503/2020\|EPI_ISL_485158\|2 | .................... | .................... | ..................... | .................... | .................... | .................... |
| E | 1844 hCoV-19/USA/LA-GBCL-26556509/2020\|EPI_ISL_485161\|2 | .................... | .................... | ..................... | .................... | .................... | .................... |
| E | 1845 hCoV-19/USA/LA-GBCL-26556517/2020\|EPI_ISL_485164\|2 | .................... | .................... | ..................... | .................... | .................... | .................... |
| E | 1846 hCoV-19/USA/LA-GBCL-26556519/2020\|EPI_ISL_485165\|2 | .................... | .................... | ..................... | .................... | .................... | .................... |
| E | 1847 hCoV-19/USA/LA-GBCL-26556523/2020\|EPI_ISL_485167\|2 | .................... | .................... | ..................... | .................... | .................... | .................... |
| E | 1848 hCoV-19/USA/LA-GBCL-26556525/2020\|EPI_ISL_485169\|2 | .................... | .................... | ..................... | .................... | .................... | .................... |
| E | 1849 hCoV-19/USA/LA-GBCL-26556526/2020\|EPI_ISL_485170\|2 | .................... | .................... | ..................... | .................... | .................... | .................... |
| E | 1850 hCoV-19/USA/LA-GBCL-26556534/2020\|EPI_ISL_485172\|2 | .................... | .................... | ..................... | .................... | .................... | .................... |
| E | 1851 hCoV-19/USA/LA-GBCL-26556535/2020\|EPI_ISL_485173\|2 | .................... | .................... | ..................... | .................... | .................... | .................... |
| E | 1852 hCoV-19/USA/LA-GBCL-26556536/2020\|EPI_ISL_485174\|2 | .................... | .................... | ..................... | .................... | .................... | .................... |
| E | 1853 hCoV-19/USA/LA-GBCL-26556539/2020\|EPI_ISL_485177\|2 | .................... | .................... | ..................... | .................... | .................... | .................... |
| E | 1854 hCoV-19/USA/LA-GBCL-26556540/2020\|EPI_ISL_485178\|2 | .................... | .................... | ..................... | .................... | .................... | .................... |
| E | 1855 hCoV-19/USA/LA-GBCL-26556543/2020\|EPI_ISL_485181\|2 | .................... | .................... | ..................... | .................... | .................... | .................... |
| E | 1856 hCoV-19/USA/LA-GBCL-26556545/2020\|EPI_ISL_485182\|2 | .................... | .................... | ..................... | .................... | .................... | .................... |
| E | 1857 hCoV-19/USA/LA-GBCL-26556550/2020\|EPI_ISL_485185\|2 | .................... | .................... | ..................... | .................... | .................... | .................... |
| E | 1858 hCoV-19/USA/LA-GBCL-26556556/2020\|EPI_ISL_485186\|2 | .................... | .................... | ..................... | .................... | .................... | .................... |
| E | 1859 hCoV-19/USA/LA-GBCL-26556557/2020\|EPI_ISL_485187\|2 | .................... | .................... | ..................... | .................... | .................... | .................... |
| E | 1860 hCoV-19/India/KA-nimh-10559/2020\|EPI_ISL_486400\|20 | .................... | .................... | ..................... | .................... | .................... | .................... |
| E | 1861 hCoV-19/India/KA-nimh-14709/2020\|EPI_ISL_486405\|20 | .................... | .................... | ..................... | .................... | .................... | .................... |
| E | 1862 hCoV-19/India/KA-nimh-15819/2020\|EPI_ISL_486408\|20 | .................... | .................... | ..................... | .................... | .................... | .................... |
| E | 1863 hCoV-19/India/KA-nimh-15896/2020\|EPI_ISL_486409\|20 | .................... | .................... | ..................... | .................... | .................... | .................... |
| E | 1864 hCoV-19/Latvia/027/2020\|EPI_ISL_486410\|2020-05-09 | .................... | .................... | ..................... | .................... | .................... | .................... |
| E | 1865 hCoV-19/Latvia/028/2020\|EPI_ISL_486411\|2020-05-07 | .................... | .................... | ..................... | .................... | .................... | .................... |
| E | 1866 hCoV-19/Latvia/029/2020\|EPI_ISL_486412\|2020-04-29 | .................... | .................... | ..................... | .................... | .................... | .................... |
| E | 1867 hCoV-19/Latvia/030/2020\|EPI_ISL_486413\|2020-05-10 | .................... | .................... | ..................... | .................... | .................... | .................... |
| E | 1868 hCoV-19/Latvia/031/2020\|EPI_ISL_486414\|2020-05-07 | .................... | .................... | ..................... | .................... | .................... | .................... |
| E | 1869 hCoV-19/Latvia/032/2020\|EPI_ISL_486415\|2020-04-27 | .................... | ....G............... | ..................... | .................... | .................... | .................... |
| E | 1870 hCoV-19/Latvia/033/2020\|EPI_ISL_486416\|2020-04-27 | .................... | .................... | ..................... | .................... | .................... | .................... |
| E | 1871 hCoV-19/Latvia/034/2020\|EPI_ISL_486417\|2020-05-08 | .................... | .................... | ..................... | .................... | .................... | .................... |
| E | 1872 hCoV-19/Latvia/035/2020\|EPI_ISL_486418\|2020-05-09 | .................... | .................... | ..................... | .................... | .................... | .................... |
| E | 1873 hCoV-19/Latvia/036/2020\|EPI_ISL_486419\|2020-04-29 | .................... | .................... | ..................... | .................... | .................... | .................... |
| E | 1874 hCoV-19/Latvia/037/2020\|EPI_ISL_486420\|2020-04-29 | .................... | .................... | ..................... | .................... | .................... | .................... |
| E | 1875 hCoV-19/Latvia/038/2020\|EPI_ISL_486421\|2020-04-27 | .................... | .................... | ..................... | .................... | .................... | .................... |
| E | 1876 hCoV-19/Latvia/039/2020\|EPI_ISL_486422\|2020-03-15 | .................... | .................... | ..................... | .................... | .................... | .................... |
| E | 1877 hCoV-19/Latvia/040/2020\|EPI_ISL_486423\|2020-03-17 | .................... | .................... | ..................... | .................... | .................... | .................... |
| E | 1878 hCoV-19/Latvia/041/2020\|EPI_ISL_486424\|2020-03-22 | .................... | .................... | ..................... | .................... | .................... | .................... |
| E | 1879 hCoV-19/Latvia/042/2020\|EPI_ISL_486425\|2020-03-22 | .................... | .................... | ..................... | .................... | .................... | .................... |
| E | 1880 hCoV-19/Latvia/044/2020\|EPI_ISL_486426\|2020-03-22 | .................... | .................... | ..................... | .................... | .................... | .................... |
| E | 1881 hCoV-19/Brazil/UN-HIAE-SP03/2020\|EPI_ISL_486427\|20 | .................... | .................... | ..................... | .................... | .................... | .................... |
| E | 1882 hCoV-19/Latvia/045/2020\|EPI_ISL_486428\|2020-03-22 | .................... | .A.................. | ..................... | .................... | .................... | .................... |
| E | 1883 hCoV-19/Brazil/UN-HIAE-SP04/2020\|EPI_ISL_486429\|20 | .................... | .................... | ..................... | .................... | .................... | .................... |
| E | 1884 hCoV-19/Latvia/046/2020\|EPI_ISL_486430\|2020-03-22 | .................... | .................... | ..................... | .................... | .................... | .................... |
| E | 1885 hCoV-19/Latvia/047/2020\|EPI_ISL_486431\|2020-03-16 | .................... | .................... | ..................... | .................... | .................... | .................... |

| | | | | | | | | | | | |
|---|---|---|---|---|---|---|---|---|---|---|---|
| E | 1886 | hCoV-19/Latvia/048/2020 | EPI_ISL_486432 | 2020-03-16 | .................... | .................... | ..................... | .................... | .................... | .................... | |
| E | 1887 | hCoV-19/Latvia/049/2020 | EPI_ISL_486433 | 2020-03-16 | .................... | .................... | ..................... | .................... | .................... | .................... | |
| E | 1888 | hCoV-19/Latvia/050/2020 | EPI_ISL_486434 | 2020-03-16 | .................... | .................... | ..................... | .................... | .................... | .................... | |
| E | 1889 | hCoV-19/Latvia/051/2020 | EPI_ISL_486435 | 2020-03-23 | .................... | .................... | ..................... | .................... | .................... | .................... | |
| E | 1890 | hCoV-19/Latvia/052/2020 | EPI_ISL_486436 | 2020-03-25 | .................... | .................... | ..................... | .................... | .................... | .................... | |
| E | 1891 | hCoV-19/Latvia/053/2020 | EPI_ISL_486437 | 2020-04-30 | .................... | .................... | ..................... | .................... | .................... | .................... | |
| E | 1892 | hCoV-19/Latvia/054/2020 | EPI_ISL_486438 | 2020-05-28 | .................... | .................... | ..................... | .................... | .................... | .................... | |
| E | 1895 | hCoV-19/Switzerland/VD-ETHZ-110008/2020 | EPI_ISL_48 | | .................... | .................... | ..................... | .................... | .................... | .................... | |
| E | 1917 | hCoV-19/Switzerland/GR-ETHZ-110049/2020 | EPI_ISL_48 | | .................... | .................... | ..................... | .................... | .......S............ | .................... | |
| E | 1922 | hCoV-19/Switzerland/TI-ETHZ-110061/2020 | EPI_ISL_48 | | .................... | .................... | ..................... | .................... | .......S............ | .................... | |
| E | 1928 | hCoV-19/Switzerland/AG-ETHZ-110070/2020 | EPI_ISL_48 | | .................... | .................... | ..................... | .................... | .................... | .................... | |
| E | 1929 | hCoV-19/Scotland/EDB3373/2020 | EPI_ISL_487700 | 2020- | .................... | ...................A | ..................... | .................... | .................... | .................... | 1 |
| E | 1930 | hCoV-19/Scotland/EDB3401/2020 | EPI_ISL_487702 | 2020- | .................... | ...................A | ..................... | .................... | .................... | .................... | 1 |
| E | 1931 | hCoV-19/Scotland/EDB2894/2020 | EPI_ISL_487703 | 2020- | .................... | .................... | ..................... | .................... | .................... | .................... | |
| E | 1932 | hCoV-19/Scotland/EDB6409/2020 | EPI_ISL_487704 | 2020- | .................... | .................... | ..................... | .................... | .................... | .................... | |
| E | 1933 | hCoV-19/Scotland/EDB5528/2020 | EPI_ISL_487705 | 2020- | .................... | ...................A | ..................... | .................... | .................... | .................... | 1 |
| E | 1934 | hCoV-19/Scotland/EDB3355/2020 | EPI_ISL_487706 | 2020- | .................... | .A.................. | ..................... | .................... | .................... | .................... | |
| E | 1935 | hCoV-19/Scotland/EDB5554/2020 | EPI_ISL_487707 | 2020- | .................... | .................... | ..................... | .................... | .................... | .................... | |
| E | 1936 | hCoV-19/Scotland/EDB3360/2020 | EPI_ISL_487708 | 2020- | .................... | ...................A | ..................... | .................... | .................... | .................... | 1 |
| E | 1937 | hCoV-19/Scotland/EDB3180/2020 | EPI_ISL_487709 | 2020- | .................... | .A.................. | ..................... | .................... | .................... | .................... | |
| E | 1938 | hCoV-19/Scotland/EDB5663/2020 | EPI_ISL_487710 | 2020- | .................... | .................... | ..................... | .................... | .................... | .................... | |
| E | 1939 | hCoV-19/Scotland/EDB6402/2020 | EPI_ISL_487711 | 2020- | .................... | .................... | ..................... | .................... | .................... | .................... | |
| E | 1940 | hCoV-19/Scotland/EDB3350/2020 | EPI_ISL_487712 | 2020- | .................... | ...................A | ..................... | .................... | .................... | .................... | 1 |
| E | 1941 | hCoV-19/Scotland/EDB6313/2020 | EPI_ISL_487713 | 2020- | .................... | .............A...... | ..................... | .................... | .................... | .................... | |
| E | 1942 | hCoV-19/Scotland/EDB3534/2020 | EPI_ISL_487715 | 2020- | .................... | .................... | ..................... | .................... | .................... | .................... | |
| E | 1943 | hCoV-19/Scotland/EDB6276/2020 | EPI_ISL_487716 | 2020- | .................... | ...................A | ..................... | .................... | .................... | .................... | 1 |
| E | 1944 | hCoV-19/Scotland/EDB5415/2020 | EPI_ISL_487717 | 2020- | .................... | ...................A | ..................... | .................... | .................... | .................... | 1 |
| E | 1945 | hCoV-19/Scotland/EDB6266/2020 | EPI_ISL_487718 | 2020- | .................... | .................... | ..................... | .................... | .................... | .................... | |
| E | 1946 | hCoV-19/Scotland/EDB3215/2020 | EPI_ISL_487719 | 2020- | .................... | .................... | ..................... | .................... | .................... | .................... | |
| E | 1947 | hCoV-19/Scotland/EDB6269/2020 | EPI_ISL_487720 | 2020- | .................... | ...................A | ..................... | .................... | .................... | .................... | 1 |
| E | 1948 | hCoV-19/Scotland/EDB5543/2020 | EPI_ISL_487721 | 2020- | .................... | .................... | ..................... | .................... | .................... | .................... | |
| E | 1949 | hCoV-19/Scotland/EDB3549/2020 | EPI_ISL_487722 | 2020- | .................... | ...................A | ..................... | .................... | .................... | .................... | 1 |
| E | 1950 | hCoV-19/Scotland/EDB5514/2020 | EPI_ISL_487723 | 2020- | .................... | .................... | ..................... | .................... | .................... | .................... | |
| E | 1951 | hCoV-19/Scotland/EDB6288/2020 | EPI_ISL_487724 | 2020- | .................... | .................... | ..................... | .................... | .................... | .................... | |
| E | 1952 | hCoV-19/Scotland/EDB3414/2020 | EPI_ISL_487725 | 2020- | .................... | .A.................. | ..................... | .................... | .................... | .................... | |
| E | 1953 | hCoV-19/Scotland/EDB3429/2020 | EPI_ISL_487726 | 2020- | .................... | ...................A | ..................... | .................... | .................... | .................... | 1 |
| E | 1954 | hCoV-19/Scotland/EDB3325/2020 | EPI_ISL_487727 | 2020- | .................... | ...................A | ..................... | .................... | .................... | .................... | 1 |
| E | 1955 | hCoV-19/Scotland/EDB2723/2020 | EPI_ISL_487728 | 2020- | .................... | .................... | ..................... | .................... | .................... | .................... | |
| E | 1956 | hCoV-19/Scotland/EDB6418/2020 | EPI_ISL_487729 | 2020- | .................... | .................... | ..................... | .................... | .................... | .................... | |
| E | 1957 | hCoV-19/Scotland/EDB3389/2020 | EPI_ISL_487730 | 2020- | .................... | ...................A | ..................... | .................... | .................... | .................... | 1 |
| E | 1958 | hCoV-19/Scotland/EDB6009/2020 | EPI_ISL_487731 | 2020- | .................... | .................... | ..................... | .................... | .................... | .................... | |
| E | 1959 | hCoV-19/Scotland/EDB5988/2020 | EPI_ISL_487732 | 2020- | .................... | .A.................. | ..................... | .................... | .................... | .................... | |
| E | 1960 | hCoV-19/Scotland/EDB2983/2020 | EPI_ISL_487733 | 2020- | .................... | .................... | ..................... | .................... | .................... | .................... | |
| E | 1961 | hCoV-19/Scotland/EDB5965/2020 | EPI_ISL_487734 | 2020- | .................... | .A.................. | ..................... | .................... | .................... | .................... | |
| E | 1962 | hCoV-19/Scotland/EDB4731/2020 | EPI_ISL_487735 | 2020- | .................... | .............A...... | ..................... | .................... | .................... | .................... | |
| E | 1963 | hCoV-19/Scotland/EDB5823/2020 | EPI_ISL_487736 | 2020- | .................... | .A.................. | ..................... | .................... | .................... | .................... | |
| E | 1964 | hCoV-19/Scotland/EDB5836/2020 | EPI_ISL_487737 | 2020- | .................... | ...................A | ..................... | .................... | .................... | .................... | 1 |
| E | 1965 | hCoV-19/Scotland/EDB5065/2020 | EPI_ISL_487739 | 2020- | .................... | .................... | ..................... | .................... | .................... | .................... | |
| E | 1966 | hCoV-19/Scotland/EDB3064/2020 | EPI_ISL_487740 | 2020- | .................... | .................... | ..................... | .................... | .................... | .................... | |
| E | 1967 | hCoV-19/Scotland/EDB3367/2020 | EPI_ISL_487741 | 2020- | .................... | .................... | ..................... | .................... | .................... | .................... | |
| E | 1968 | hCoV-19/Scotland/EDB3399/2020 | EPI_ISL_487742 | 2020- | .................... | ...................A | ..................... | .................... | .................... | .................... | 1 |
| E | 1969 | hCoV-19/Scotland/EDB3354/2020 | EPI_ISL_487743 | 2020- | .................... | .................... | ..................... | .................... | .................... | .................... | |
| E | 1970 | hCoV-19/Scotland/EDB5428/2020 | EPI_ISL_487745 | 2020- | .................... | .................... | ..................... | .................... | .................... | .................... | |
| E | 1971 | hCoV-19/Scotland/EDB3358/2020 | EPI_ISL_487746 | 2020- | .................... | .................... | ..................... | .................... | .................... | .................... | |
| E | 1972 | hCoV-19/Scotland/EDB3512/2020 | EPI_ISL_487747 | 2020- | .................... | .A.................. | ..................... | .................... | .................... | .................... | |
| E | 1973 | hCoV-19/Scotland/EDB2399/2020 | EPI_ISL_487749 | 2020- | .................... | .................... | ..................... | .................... | .................... | .................... | |
| E | 1974 | hCoV-19/Scotland/EDB3517/2020 | EPI_ISL_487750 | 2020- | .................... | .................... | ..................... | .................... | .................... | .................... | |
| E | 1975 | hCoV-19/Scotland/EDB5953/2020 | EPI_ISL_487751 | 2020- | .................... | ...................A | ..................... | .................... | .................... | .................... | 1 |
| E | 1976 | hCoV-19/Scotland/EDB4276/2020 | EPI_ISL_487752 | 2020- | .................... | ...................A | ..................... | .................... | .................... | .................... | 1 |
| E | 1977 | hCoV-19/Scotland/EDB3526/2020 | EPI_ISL_487753 | 2020- | .................... | .................... | ..................... | .................... | .................... | .................... | |
| E | 1978 | hCoV-19/Scotland/EDB5489/2020 | EPI_ISL_487754 | 2020- | .................... | .................... | ..................... | .................... | .................... | .................... | |
| E | 1979 | hCoV-19/Scotland/EDB5512/2020 | EPI_ISL_487755 | 2020- | .................... | .................... | ..................... | .................... | .................... | .................... | |
| E | 1980 | hCoV-19/Scotland/EDB5503/2020 | EPI_ISL_487756 | 2020- | .................... | .................... | ..................... | .................... | .................... | .................... | |
| E | 1981 | hCoV-19/Scotland/EDB3397/2020 | EPI_ISL_487757 | 2020- | .................... | .................... | ..................... | .................... | .................... | .................... | |
| E | 1982 | hCoV-19/Scotland/EDB5879/2020 | EPI_ISL_487758 | 2020- | .................... | ...................A | ..................... | .................... | .................... | .................... | 1 |
| E | 1983 | hCoV-19/Scotland/EDB3445/2020 | EPI_ISL_487759 | 2020- | .................... | .................... | ..................... | .................... | .................... | .................... | |
| E | 1984 | hCoV-19/Scotland/EDB4331/2020 | EPI_ISL_487760 | 2020- | .................... | .A.................. | ..................... | .................... | .................... | .................... | |
| E | 1985 | hCoV-19/Scotland/EDB5506/2020 | EPI_ISL_487761 | 2020- | .................... | .................... | ..................... | .................... | .................... | .................... | |
| E | 1986 | hCoV-19/Scotland/EDB5405/2020 | EPI_ISL_487762 | 2020- | .................... | .................... | ..................... | .................... | .................... | .................... | |
| E | 1987 | hCoV-19/Scotland/EDB5803/2020 | EPI_ISL_487763 | 2020- | .................... | ...................A | ..................... | .................... | .................... | .................... | 1 |
| E | 1988 | hCoV-19/Scotland/EDB2721/2020 | EPI_ISL_487764 | 2020- | .................... | .................... | ..................... | .................... | .................... | .................... | |
| E | 1989 | hCoV-19/Scotland/EDB6410/2020 | EPI_ISL_487765 | 2020- | .................... | .................... | ..................... | .................... | .................... | .................... | |
| E | 1990 | hCoV-19/Scotland/EDB5795/2020 | EPI_ISL_487766 | 2020- | .................... | ...................A | ..................... | .................... | .................... | .................... | 1 |
| E | 1991 | hCoV-19/Scotland/EDB5825/2020 | EPI_ISL_487768 | 2020- | .................... | ...................A | ..................... | .................... | .................... | .................... | 1 |
| E | 1992 | hCoV-19/Scotland/EDB3527/2020 | EPI_ISL_487769 | 2020- | .................... | .................... | ..................... | .................... | .................... | .................... | |
| E | 1993 | hCoV-19/Scotland/EDB3330/2020 | EPI_ISL_487770 | 2020- | .................... | .A.................. | ..................... | .................... | .................... | .................... | |
| E | 1994 | hCoV-19/Scotland/EDB3412/2020 | EPI_ISL_487771 | 2020- | .................... | ...................A | ..................... | .................... | .................... | .................... | 1 |
| E | 1995 | hCoV-19/Scotland/EDB5777/2020 | EPI_ISL_487772 | 2020- | .................... | .................... | ..................... | .................... | .................... | .................... | |
| E | 1996 | hCoV-19/Scotland/EDB5964/2020 | EPI_ISL_487774 | 2020- | .................... | .................... | ..................... | .................... | .................... | .................... | |
| E | 1997 | hCoV-19/Scotland/EDB5862/2020 | EPI_ISL_487775 | 2020- | .................... | .................... | ..................... | .................... | .................... | .................... | |

| | | | | | | | | |
|---|---|---|---|---|---|---|---|---|
| E | 1998 hCoV-19/Scotland/EDB6023/2020\|EPI_ISL_487776\|2020- | ···················· | ·A·················· | ···················· | ···················· | ···················· | ···················· | |
| E | 1999 hCoV-19/Scotland/EDB5888/2020\|EPI_ISL_487777\|2020- | ···················· | ···················A | ···················· | ···················· | ···················· | ···················· | 1 |
| E | 2000 hCoV-19/Scotland/EDB4222/2020\|EPI_ISL_487779\|2020- | ···················· | ···················A | ···················· | ···················· | ···················· | ···················· | 1 |
| E | 2001 hCoV-19/Scotland/EDB3572/2020\|EPI_ISL_487780\|2020- | ···················· | ···················· | ···················· | ···················· | ···················· | ···················· | |
| E | 2002 hCoV-19/Scotland/EDB5089/2020\|EPI_ISL_487781\|2020- | ···················· | ···················· | ···················· | ···················· | ···················· | ···················· | |
| E | 2003 hCoV-19/Scotland/EDB3396/2020\|EPI_ISL_487782\|2020- | ···················· | ···················A | ···················· | ···················· | ···················· | ···················· | 1 |
| E | 2004 hCoV-19/Scotland/EDB6052/2020\|EPI_ISL_487783\|2020- | ···················· | ···················· | ···················· | ···················· | ···················· | ···················· | |
| E | 2005 hCoV-19/Scotland/EDB6545/2020\|EPI_ISL_487784\|2020- | ···················· | ···················· | ···················· | ···················· | ···················· | ···················· | |
| E | 2006 hCoV-19/Scotland/EDB5827/2020\|EPI_ISL_487786\|2020- | ···················· | ···················A | ···················· | ···················· | ···················· | ···················· | 1 |
| E | 2007 hCoV-19/Scotland/EDB5998/2020\|EPI_ISL_487787\|2020- | ···················· | ···················A | ···················· | ···················· | ···················· | ···················· | 1 |
| E | 2008 hCoV-19/Scotland/EDB2744/2020\|EPI_ISL_487788\|2020- | ···················· | ···················· | ···················· | ···················· | ···················· | ···················· | |
| E | 2009 hCoV-19/Scotland/EDB5584/2020\|EPI_ISL_487789\|2020- | ···················· | ···················· | ···················· | ···················· | ···················· | ···················· | |
| E | 2010 hCoV-19/Scotland/EDB3371/2020\|EPI_ISL_487790\|2020- | ···················· | ···················· | ···················· | ···················· | ···················· | ···················· | |
| E | 2011 hCoV-19/Scotland/EDB5411/2020\|EPI_ISL_487791\|2020- | ···················· | ···················A | ···················· | ···················· | ···················· | ···················· | 1 |
| E | 2012 hCoV-19/Scotland/EDB6412/2020\|EPI_ISL_487792\|2020- | ···················· | ···················· | ···················· | ···················· | ···················· | ···················· | |
| E | 2013 hCoV-19/Scotland/EDB5819/2020\|EPI_ISL_487793\|2020- | ···················· | ·A·················· | ···················· | ···················· | ···················· | ···················· | |
| E | 2014 hCoV-19/Scotland/EDB2743/2020\|EPI_ISL_487794\|2020- | ···················· | ···················· | ···················· | ···················· | ···················· | ···················· | |
| E | 2015 hCoV-19/Scotland/EDB6032/2020\|EPI_ISL_487795\|2020- | ···················· | ···················· | ···················· | ···················· | ···················· | ···················· | |
| E | 2016 hCoV-19/Scotland/EDB4094/2020\|EPI_ISL_487796\|2020- | ···················· | ·A·················· | ···················· | ···················· | ···················· | ···················· | |
| E | 2017 hCoV-19/Scotland/EDB6435/2020\|EPI_ISL_487797\|2020- | ···················· | ···················A | ···················· | ···················· | ···················· | ···················· | 1 |
| E | 2018 hCoV-19/Scotland/EDB5980/2020\|EPI_ISL_487798\|2020- | ···················· | ···················A | ···················· | ···················· | ···················· | ···················· | 1 |
| E | 2019 hCoV-19/Scotland/EDB5511/2020\|EPI_ISL_487799\|2020- | ···················· | ···················· | ···················· | ···················· | ···················· | ···················· | |
| E | 2020 hCoV-19/England/BRIS-18532FE/2020\|EPI_ISL_492242\|2 | ···················· | ···················· | ···················· | ···················· | ···················· | ···················· | |
| E | 2021 hCoV-19/England/BRIS-18539DB/2020\|EPI_ISL_492243\|2 | ···················· | ···················· | ···················· | ···················· | ···················· | ···················· | |
| E | 2022 hCoV-19/England/BRIS-1853CB4/2020\|EPI_ISL_492244\|2 | ···················· | ···A················ | ···················· | ···················· | ···················· | ···················· | |
| E | 2023 hCoV-19/England/BRIS-1853B6C/2020\|EPI_ISL_492246\|2 | ···················· | ···················· | ···················· | ···················· | ···················· | ···················· | |
| E | 2024 hCoV-19/England/BRIS-1854257/2020\|EPI_ISL_492249\|2 | ···················· | ···················· | ···················· | ···················· | ···················· | ···················· | |
| E | 2025 hCoV-19/England/BRIS-185414B/2020\|EPI_ISL_492250\|2 | ···················· | ···················· | ···················· | ···················· | ···················· | ···················· | |
| E | 2026 hCoV-19/England/BRIS-18542EE/2020\|EPI_ISL_492274\|2 | ···················· | ···················· | ···················· | ···················· | ···················· | ···················· | |
| E | 2027 hCoV-19/England/BRIS-1853F7F/2020\|EPI_ISL_492277\|2 | ···················· | ···················· | ···················· | ···················· | ···················· | ···················· | |
| E | 2028 hCoV-19/England/BRIS-18541F0/2020\|EPI_ISL_492278\|2 | ···················· | ···················· | ···················· | ···················· | ···················· | ···················· | |
| E | 2029 hCoV-19/England/BRIS-1853A9C/2020\|EPI_ISL_492279\|2 | ···················· | ···················· | ···················· | ···················· | ···················· | ···················· | |
| E | 2030 hCoV-19/England/BRIS-18536B6/2020\|EPI_ISL_492281\|2 | ···················· | ···················· | ···················· | ···················· | ···················· | ···················· | |
| E | 2031 hCoV-19/England/BRIS-1854345/2020\|EPI_ISL_492283\|2 | ···················· | ···················· | ···················· | ···················· | ···················· | ···················· | |
| E | 2032 hCoV-19/England/BRIS-1853B4E/2020\|EPI_ISL_492284\|2 | ···················· | ···················· | ···················· | ···················· | ···················· | ···················· | |
| E | 2033 hCoV-19/England/BRIS-18530B8/2020\|EPI_ISL_492285\|2 | ···················· | ···················· | ···················· | ···················· | ···················· | ···················· | |
| E | 2034 hCoV-19/England/BRIS-1853249/2020\|EPI_ISL_492286\|2 | ···················· | ···················· | ···················· | ···················· | ···················· | ···················· | |
| E | 2035 hCoV-19/England/BRIS-18534DA/2020\|EPI_ISL_492287\|2 | ···················· | ···················· | ···················· | ···················· | ···················· | ···················· | |
| E | 2036 hCoV-19/England/BRIS-1853DEE/2020\|EPI_ISL_492289\|2 | ···················· | ···················· | ···················· | ···················· | ···················· | ···················· | |
| E | 2037 hCoV-19/England/BRIS-185362F/2020\|EPI_ISL_492290\|2 | ···················· | ···················· | ···················· | ···················· | ···················· | ···················· | |
| E | 2038 hCoV-19/England/BRIS-1853197/2020\|EPI_ISL_492291\|2 | ···················· | ···················· | ···················· | ···················· | ···················· | ···················· | |
| E | 2039 hCoV-19/England/BRIS-18540A8/2020\|EPI_ISL_492292\|2 | ···················· | ···················· | ···················· | ···················· | ···················· | ···················· | |
| E | 2040 hCoV-19/England/BRIS-1854002/2020\|EPI_ISL_492294\|2 | ···················· | ···················· | ···················· | ···················· | ···················· | ···················· | |
| E | 2041 hCoV-19/England/BRIS-1852F25/2020\|EPI_ISL_492298\|2 | ···················· | ···················· | ···················· | ···················· | ···················· | ···················· | |
| E | 2042 hCoV-19/England/BRIS-1853F42/2020\|EPI_ISL_492299\|2 | ···················· | ···················· | ···················· | ···················· | ···················· | ···················· | |
| E | 2043 hCoV-19/England/NOTT-1129E1/2020\|EPI_ISL_493572\|20 | ···················· | ···················· | ···················· | ···················· | ···················· | ···················· | |
| E | 2044 hCoV-19/England/NOTT-112A1B/2020\|EPI_ISL_493574\|20 | ···················· | ···················· | ···················· | ···················· | ···················· | ···················· | |
| E | 2045 hCoV-19/England/NOTT-112A39/2020\|EPI_ISL_493576\|20 | ···················· | ···················· | ···················· | ···················· | ···················· | ···················· | |
| E | 2046 hCoV-19/England/NOTT-112A57/2020\|EPI_ISL_493578\|20 | ···················· | ···················· | ···················· | ···················· | ···················· | ···················· | |
| E | 2047 hCoV-19/England/NOTT-112A93/2020\|EPI_ISL_493580\|20 | ···················· | ···················· | ···················· | ···················· | ···················· | ···················· | |
| E | 2048 hCoV-19/England/NOTT-112AA2/2020\|EPI_ISL_493581\|20 | ···················· | ···················· | ···················· | ···················· | ···················· | ···················· | |
| E | 2049 hCoV-19/Japan/PG-0544/2020\|EPI_ISL_480200\|2020-03- | ···················· | ···················· | ···················· | ···················· | ···················· | ···················· | |
| E | 2050 hCoV-19/Japan/PG-0545/2020\|EPI_ISL_480201\|2020-03- | ···················· | ···················· | ···················· | ···················· | ···················· | ···················· | |
| E | 2051 hCoV-19/Japan/PG-0546/2020\|EPI_ISL_480202\|2020-04- | ···················· | ···················· | ···················· | ···················· | ···················· | ···················· | |
| E | 2052 hCoV-19/Japan/PG-0547/2020\|EPI_ISL_480203\|2020-04- | ···················· | ···················· | ···················· | ···················· | ···················· | ···················· | |
| E | 2053 hCoV-19/Japan/PG-0560/2020\|EPI_ISL_480210\|2020-04- | ···················· | ···················· | ···················· | ···················· | ···················· | ···················· | |
| E | 2054 hCoV-19/Japan/PG-0561/2020\|EPI_ISL_480211\|2020-04- | ···················· | ···················· | ···················· | ···················· | ···················· | ···················· | |
| E | 2055 hCoV-19/Japan/PG-0563/2020\|EPI_ISL_480212\|2020-04- | ···················· | ···················· | ···················· | ···················· | ···················· | ···················· | |
| E | 2056 hCoV-19/Japan/PG-0564/2020\|EPI_ISL_480213\|2020-04- | ···················· | ···················· | ···················· | ···················· | ···················· | ···················· | |
| E | 2057 hCoV-19/Japan/PG-0566/2020\|EPI_ISL_480214\|2020-04- | ···················· | ···················· | ···················· | ···················· | ···················· | ···················· | |
| E | 2058 hCoV-19/Japan/PG-0572/2020\|EPI_ISL_480215\|2020-04- | ···················· | ···················· | ···················· | ···················· | ···················· | ···················· | |
| E | 2059 hCoV-19/Japan/PG-0573/2020\|EPI_ISL_480216\|2020-04- | ···················· | ···················· | ···················· | ···················· | ···················· | ···················· | |
| E | 2060 hCoV-19/Japan/PG-0576/2020\|EPI_ISL_480217\|2020-04- | ···················· | ···················· | ···················· | ···················· | ···················· | ···················· | |
| E | 2061 hCoV-19/Japan/PG-0579/2020\|EPI_ISL_480218\|2020-04- | ···················· | ···················· | ···················· | ···················· | ···················· | ···················· | |
| E | 2062 hCoV-19/Japan/PG-0582/2020\|EPI_ISL_480219\|2020-04- | ···················· | ···················· | ···················· | ···················· | ···················· | ···················· | |
| E | 2063 hCoV-19/Japan/PG-0587/2020\|EPI_ISL_480220\|2020-04- | ···················· | ···················· | ···················· | ···················· | ···················· | ···················· | |
| E | 2064 hCoV-19/Japan/PG-0589/2020\|EPI_ISL_480221\|2020-04- | ···················· | ···················· | ···················· | ···················· | ···················· | ···················· | |
| E | 2065 hCoV-19/Japan/PG-0590/2020\|EPI_ISL_480222\|2020-04- | ···················· | ···················· | ···················· | ···················· | ···················· | ···················· | |
| E | 2066 hCoV-19/Japan/PG-0591/2020\|EPI_ISL_480223\|2020-04- | ···················· | ···················· | ···················· | ···················· | ···················· | ···················· | |
| E | 2067 hCoV-19/Bulgaria/10/2020\|EPI_ISL_480224\|2020-04-29 | ···················· | ···················· | ···················· | ···················· | ···················· | ···················· | |
| E | 2068 hCoV-19/Turkey/IST-GLAB-003/2020\|EPI_ISL_480228\|20 | ···················· | ···················· | ···················· | ···················· | ···················· | ···················· | |
| E | 2069 hCoV-19/Turkey/IST-GLAB-004/2020\|EPI_ISL_480229\|20 | ···················· | ···················· | ···················· | ···················· | ···················· | ···················· | |
| E | 2070 hCoV-19/Turkey/IST-GLAB-006/2020\|EPI_ISL_480230\|20 | ···················· | ···················· | ···················· | ···················· | ···················· | ···················· | |
| E | 2071 hCoV-19/Turkey/IST-GLAB-014/2020\|EPI_ISL_480232\|20 | ···················· | ···················· | ···················· | ···················· | ···················· | ···················· | |
| E | 2072 hCoV-19/Turkey/IST-GLAB-021/2020\|EPI_ISL_480233\|20 | ···················· | ···················· | ···················· | ···················· | ···················· | ···················· | |
| E | 2073 hCoV-19/Turkey/IST-GLAB-030/2020\|EPI_ISL_480234\|20 | ···················· | ···················· | ···················· | ···················· | ···················· | ···················· | |
| E | 2074 hCoV-19/Turkey/IST-GLAB-054/2020\|EPI_ISL_480235\|20 | ···················· | ···················· | ···················· | ···················· | ···················· | ···················· | |
| E | 2075 hCoV-19/Turkey/IST-GLAB-055/2020\|EPI_ISL_480236\|20 | ···················· | ···················· | ···················· | ···················· | ···················· | ···················· | |
| E | 2076 hCoV-19/Turkey/IST-GLAB-059/2020\|EPI_ISL_480237\|20 | ···················· | ···················· | ···················· | ···················· | ···················· | ···················· | |
| E | 2077 hCoV-19/Turkey/IST-GLAB-062/2020\|EPI_ISL_480238\|20 | ···················· | ···················· | ···················· | ···················· | ···················· | ···················· | |

| | | | | | | | |
|---|---|---|---|---|---|---|---|
| E | 2078 hCoV-19/Turkey/IST-GLAB-063/2020\|EPI_ISL_480239\|20 | ···················· | ·A·················· | ····················· | ···················· | ···················· | ··A················· |
| E | 2079 hCoV-19/Turkey/IST-GLAB-064/2020\|EPI_ISL_480240\|20 | ···················· | ···················· | ····················· | ···················· | ···················· | ···················· |
| E | 2080 hCoV-19/Turkey/IST-GLAB-086/2020\|EPI_ISL_480241\|20 | ···················· | ···················· | ····················· | ···················· | ···················· | ···················· |
| E | 2081 hCoV-19/Turkey/IST-GLAB-089/2020\|EPI_ISL_480242\|20 | ···················· | ···················· | ····················· | ···················· | ···················· | ···················· |
| E | 2082 hCoV-19/Turkey/IST-GLAB-091/2020\|EPI_ISL_480243\|20 | ···················· | ···················· | ····················· | ···················· | ···················· | ···················· |
| E | 2083 hCoV-19/Turkey/IST-GLAB-092/2020\|EPI_ISL_480244\|20 | ···················· | ···················· | ····················· | ···················· | ···················· | ···················· |
| E | 2084 hCoV-19/Turkey/IST-GLAB-094/2020\|EPI_ISL_480245\|20 | ···················· | ···················· | ····················· | ···················· | ···················· | ···················· |
| E | 2085 hCoV-19/Turkey/IST-GLAB-114/2020\|EPI_ISL_480246\|20 | ···················· | ···················· | ····················· | ···················· | ···················· | ···················· |
| E | 2086 hCoV-19/Turkey/IST-GLAB-115/2020\|EPI_ISL_480247\|20 | ···················· | ···················· | ····················· | ···················· | ···················· | ···················· |
| E | 2087 hCoV-19/Turkey/IST-GLAB-126/2020\|EPI_ISL_480248\|20 | ···················· | ···················· | ····················· | ···················· | ···················· | ···················· |
| E | 2088 hCoV-19/Turkey/IST-GLAB-139/2020\|EPI_ISL_480249\|20 | ···················· | ···················· | ····················· | ···················· | ···················· | ···················· |
| E | 2089 hCoV-19/Turkey/IST-GLAB-149/2020\|EPI_ISL_480250\|20 | ···················· | ······A············· | ····················· | ···················· | ···················· | ···················· |
| E | 2090 hCoV-19/Turkey/IST-GLAB-169/2020\|EPI_ISL_480262\|20 | ···················· | ···················· | ····················· | ···················· | ···················· | ···················· |
| E | 2091 hCoV-19/Turkey/IST-GLAB-170/2020\|EPI_ISL_480263\|20 | ···················· | ···················· | ····················· | ···················· | ···················· | ···················· |
| E | 2092 hCoV-19/Turkey/IST-GLAB-171/2020\|EPI_ISL_480264\|20 | ···················· | ···················· | ····················· | ···················· | ···················· | ···················· |
| E | 2093 hCoV-19/Turkey/IST-GLAB-173/2020\|EPI_ISL_480265\|20 | ···················· | ···················· | ····················· | ···················· | ···················· | ···················· |
| E | 2094 hCoV-19/Turkey/IST-GLAB-174/2020\|EPI_ISL_480266\|20 | ···················· | ···················· | ····················· | ···················· | ···················· | ···················· |
| E | 2095 hCoV-19/Turkey/IST-GLAB-176/2020\|EPI_ISL_480267\|20 | ···················· | ···················· | ····················· | ···················· | ···················· | ···················· |
| E | 2096 hCoV-19/Turkey/IST-GLAB-178/2020\|EPI_ISL_480268\|20 | ···················· | ···················· | ····················· | ···················· | ···················· | ···················· |
| E | 2097 hCoV-19/Turkey/IST-GLAB-179/2020\|EPI_ISL_480269\|20 | ···················· | ···················· | ····················· | ···················· | ···················· | ···················· |
| E | 2098 hCoV-19/Turkey/IST-GLAB-181/2020\|EPI_ISL_480270\|20 | ···················· | ···················· | ····················· | ···················· | ···················· | ···················· |
| E | 2099 hCoV-19/Turkey/IST-GLAB-182/2020\|EPI_ISL_480271\|20 | ···················· | ···················· | ····················· | ···················· | ···················· | ···················· |
| E | 2100 hCoV-19/Turkey/IST-GLAB-183/2020\|EPI_ISL_480272\|20 | ···················· | ···················· | ····················· | ···················· | ···················· | ···················· |
| E | 2101 hCoV-19/Turkey/IST-GLAB-185/2020\|EPI_ISL_480273\|20 | ···················· | ···················· | ····················· | ···················· | ···················· | ···················· |
| E | 2102 hCoV-19/Turkey/IST-GLAB-187/2020\|EPI_ISL_480274\|20 | ···················· | ···················· | ····················· | ···················· | ···················· | ···················· |
| E | 2103 hCoV-19/Turkey/IST-GLAB-188/2020\|EPI_ISL_480275\|20 | ···················· | ···················· | ····················· | ···················· | ···················· | ···················· |
| E | 2104 hCoV-19/Turkey/IST-GLAB-189/2020\|EPI_ISL_480276\|20 | ···················· | ···················· | ····················· | ···················· | ···················· | ···················· |
| E | 2105 hCoV-19/Turkey/IST-GLAB-190/2020\|EPI_ISL_480277\|20 | ···················· | ···················· | ····················· | ···················· | ···················· | ···················· |
| E | 2106 hCoV-19/Turkey/IST-GLAB-193/2020\|EPI_ISL_480278\|20 | ···················· | ···················· | ····················· | ···················· | ···················· | ···················· |
| E | 2107 hCoV-19/Turkey/IST-GLAB-195/2020\|EPI_ISL_480279\|20 | ···················· | ···················· | ····················· | ···················· | ···················· | ···················· |
| E | 2108 hCoV-19/Turkey/IST-GLAB-196/2020\|EPI_ISL_480280\|20 | ···················· | ···················· | ····················· | ···················· | ···················· | ···················· |
| E | 2109 hCoV-19/Turkey/IST-GLAB-197/2020\|EPI_ISL_480281\|20 | ···················· | ···················· | ····················· | ···················· | ···················· | ···················· |
| E | 2110 hCoV-19/Turkey/IST-GLAB-199/2020\|EPI_ISL_480282\|20 | ···················· | ···················· | ····················· | ···················· | ···················· | ···················· |
| E | 2111 hCoV-19/Turkey/IST-GLAB-204/2020\|EPI_ISL_480283\|20 | ···················· | ···················· | ····················· | ···················· | ···················· | ···················· |
| E | 2112 hCoV-19/Turkey/IST-GLAB-205/2020\|EPI_ISL_480284\|20 | ···················· | ···················· | ····················· | ···················· | ···················· | ···················· |
| E | 2113 hCoV-19/Turkey/IST-GLAB-206/2020\|EPI_ISL_480285\|20 | ···················· | ···················· | ····················· | ···················· | ···················· | ···················· |
| E | 2114 hCoV-19/Turkey/IST-GLAB-207/2020\|EPI_ISL_480286\|20 | ···················· | ···················· | ····················· | ···················· | ···················· | ···················· |
| E | 2115 hCoV-19/Turkey/IST-GLAB-208/2020\|EPI_ISL_480287\|20 | ···················· | ··········A········· | ····················· | ···················· | ···················· | ···················· |
| E | 2116 hCoV-19/Turkey/IST-GLAB-209/2020\|EPI_ISL_480288\|20 | ···················· | ···················· | ····················· | ···················· | ···················· | ···················· |
| E | 2117 hCoV-19/Turkey/IST-GLAB-210/2020\|EPI_ISL_480289\|20 | ···················· | ·A·················· | ····················· | ···················· | ···················· | ··A················· |
| E | 2118 hCoV-19/Turkey/IST-GLAB-216/2020\|EPI_ISL_480291\|20 | ···················· | ···················· | ····················· | ···················· | ···················· | ···················· |
| E | 2119 hCoV-19/Turkey/IST-GLAB-217/2020\|EPI_ISL_480292\|20 | ···················· | ···················· | ····················· | ···················· | ···················· | ···················· |
| E | 2120 hCoV-19/Finland/16A52S11/2020\|EPI_ISL_481513\|2020- | ···················· | ···················· | ····················· | ···················· | ···················· | ···············A···· |
| E | 2121 hCoV-19/Finland/10A10S14/2020\|EPI_ISL_481514\|2020- | ···················· | ···················· | ····················· | ···················· | ···················· | ···················· |
| E | 2122 hCoV-19/Finland/10A11S14/2020\|EPI_ISL_481515\|2020- | ···················· | ···················· | ····················· | ···················· | ···················· | ···················· |
| E | 2123 hCoV-19/Finland/10A15S14/2020\|EPI_ISL_481516\|2020- | ···················· | ···················· | ····················· | ···················· | ···················· | ···················· |
| E | 2124 hCoV-19/Finland/10A16S14/2020\|EPI_ISL_481517\|2020- | ···················· | ···················· | ····················· | ···················· | ···················· | ···················· |
| E | 2125 hCoV-19/Finland/10A17S14/2020\|EPI_ISL_481518\|2020- | ···················· | ···················· | ····················· | ···················· | ···················· | ···················· |
| E | 2126 hCoV-19/Finland/10A1S14/2020\|EPI_ISL_481519\|2020-0 | ···················· | ···················· | ····················· | ···················· | ···················· | ···················· |
| E | 2127 hCoV-19/Finland/10A20S10/2020\|EPI_ISL_481520\|2020- | ···················· | ···················· | ····················· | ···················· | ···················· | ···················· |
| E | 2128 hCoV-19/Finland/10A21S14/2020\|EPI_ISL_481521\|2020- | ···················· | ···················· | ····················· | ···················· | ···················· | ···················· |
| E | 2129 hCoV-19/Finland/10A22S14/2020\|EPI_ISL_481522\|2020- | ···················· | ···················· | ····················· | ···················· | ···················· | ···················· |
| E | 2130 hCoV-19/Finland/10A30S10/2020\|EPI_ISL_481523\|2020- | ···················· | ···················· | ····················· | ···················· | ···················· | ···················· |
| E | 2131 hCoV-19/Finland/10A30S14/2020\|EPI_ISL_481524\|2020- | ···················· | ···················· | ····················· | ···················· | ···················· | ···················· |
| E | 2132 hCoV-19/Finland/10A48S10/2020\|EPI_ISL_481525\|2020- | ···················· | ···················· | ····················· | ···················· | ···················· | ···················· |
| E | 2133 hCoV-19/Finland/10A53S14/2020\|EPI_ISL_481526\|2020- | ···················· | ···················· | ····················· | ···················· | ···················· | ···················· |
| E | 2134 hCoV-19/Finland/10A54S10/2020\|EPI_ISL_481527\|2020- | ···················· | ···················· | ····················· | ···················· | ···················· | ···················· |
| E | 2135 hCoV-19/Finland/10A57S10/2020\|EPI_ISL_481528\|2020- | ···················· | ···················· | ····················· | ···················· | ···················· | ···················· |
| E | 2136 hCoV-19/Finland/10A63S14/2020\|EPI_ISL_481529\|2020- | ···················· | ···················· | ····················· | ···················· | ···················· | ···················· |
| E | 2137 hCoV-19/Finland/10A66S14/2020\|EPI_ISL_481530\|2020- | ···················· | ···················· | ····················· | ···················· | ···················· | ···················· |
| E | 2138 hCoV-19/Finland/10A80S14/2020\|EPI_ISL_481531\|2020- | ···················· | ···················· | ····················· | ···················· | ···················· | ···················· |
| E | 2139 hCoV-19/Finland/11A27S7/2020\|EPI_ISL_481532\|2020-0 | ···················· | ···················· | ····················· | ···················· | ···················· | ···················· |
| E | 2140 hCoV-19/Finland/11A36S7/2020\|EPI_ISL_481533\|2020-0 | ···················· | ···················· | ····················· | ···················· | ···················· | ···················· |
| E | 2141 hCoV-19/Finland/11A39S7/2020\|EPI_ISL_481534\|2020-0 | ···················· | ···················· | ····················· | ···················· | ···················· | ···················· |
| E | 2142 hCoV-19/Finland/12A46SVI/2020\|EPI_ISL_481540\|2020- | ···················· | ···················· | ····················· | ···················· | ···················· | ···················· |
| E | 2143 hCoV-19/Finland/12A47SVI/2020\|EPI_ISL_481541\|2020- | ···················· | ···················· | ····················· | ···················· | ···················· | ···················· |
| E | 2144 hCoV-19/Finland/12A49SVI/2020\|EPI_ISL_481542\|2020- | ···················· | ···················· | ····················· | ···················· | ···················· | ···················· |
| E | 2145 hCoV-19/Finland/12A53SVI/2020\|EPI_ISL_481543\|2020- | ···················· | ···················· | ····················· | ···················· | ···················· | ···················· |
| E | 2146 hCoV-19/Finland/12A55SVI/2020\|EPI_ISL_481544\|2020- | ···················· | ···················· | ····················· | ···················· | ···················· | ···················· |
| E | 2147 hCoV-19/Finland/12A55SVIIB/2020\|EPI_ISL_481545\|202 | ···················· | ···················· | ····················· | ···················· | ···················· | ···················· |
| E | 2148 hCoV-19/Finland/12A63SVI/2020\|EPI_ISL_481546\|2020- | ···················· | ···················· | ····················· | ···················· | ···················· | ···················· |
| E | 2149 hCoV-19/Finland/12A67SVIIC/2020\|EPI_ISL_481547\|202 | ···················· | ···················· | ····················· | ···················· | ···················· | ···················· |
| E | 2150 hCoV-19/Finland/12A70SVIIC/2020\|EPI_ISL_481548\|202 | ···················· | ···················· | ····················· | ···················· | ···················· | ···················· |
| E | 2151 hCoV-19/Finland/13M6S4/2020\|EPI_ISL_481549\|2020-03 | ···················· | ···················· | ····················· | ···················· | ···················· | ···················· |
| E | 2152 hCoV-19/Finland/13May68S20/2020\|EPI_ISL_481550\|202 | ···················· | ···················· | ····················· | ···················· | ···················· | ···················· |
| E | 2153 hCoV-19/Finland/14A16S15/2020\|EPI_ISL_481551\|2020- | ···················· | ···················· | ····················· | ···················· | ···················· | ···················· |
| E | 2154 hCoV-19/Finland/14A51S15/2020\|EPI_ISL_481552\|2020- | ···················· | ···················· | ····················· | ···················· | ···················· | ···················· |
| E | 2155 hCoV-19/Finland/14A67S15/2020\|EPI_ISL_481553\|2020- | ···················· | ···················· | ····················· | ···················· | ···················· | ···················· |
| E | 2156 hCoV-19/Finland/14A69S15/2020\|EPI_ISL_481554\|2020- | ···················· | ···················· | ····················· | ···················· | ···················· | ···················· |
| E | 2157 hCoV-19/Finland/15A32S5/2020\|EPI_ISL_481555\|2020-0 | ···················· | ···················· | ····················· | ···················· | ···················· | ···················· |

| | | | | | | | | |
|---|---|---|---|---|---|---|---|---|
| E | 2158 | hCoV-19/Finland/15A36S5/2020\|EPI_ISL_481556\|2020-0 | ···················· | ···················· | ···················· | ···················· | ···················· | ···················· |
| E | 2159 | hCoV-19/Finland/15A41S5/2020\|EPI_ISL_481557\|2020-0 | ···················· | ···················· | ···················· | ···················· | ···················· | ···················· |
| E | 2160 | hCoV-19/Finland/15A50S5/2020\|EPI_ISL_481558\|2020-0 | ···················· | ···················· | ···················· | ···················· | ···················· | ···················· |
| E | 2161 | hCoV-19/Finland/15A602/2020\|EPI_ISL_481559\|2020-04 | ···················· | ···················· | ···················· | ···················· | ···················· | ···················· |
| E | 2162 | hCoV-19/Finland/15A628/2020\|EPI_ISL_481560\|2020-04 | ···················· | ···················· | ···················· | ···················· | ···················· | ···················· |
| E | 2163 | hCoV-19/Finland/15A84S16/2020\|EPI_ISL_481561\|2020- | ···················· | ···················· | ···················· | ···················· | ···················· | ···················· |
| E | 2164 | hCoV-19/Finland/15A92S5/2020\|EPI_ISL_481562\|2020-0 | ···················· | ···················· | ···················· | ···················· | ···················· | ···················· |
| E | 2165 | hCoV-19/Finland/15A93S5/2020\|EPI_ISL_481563\|2020-0 | ···················· | ···················· | ···················· | ···················· | ···················· | ···················· |
| E | 2166 | hCoV-19/Finland/15A99S16/2020\|EPI_ISL_481564\|2020- | ···················· | ···················· | ···················· | ···················· | ···················· | ···················· |
| E | 2167 | hCoV-19/Finland/15MAY16S13/2020\|EPI_ISL_481565\|202 | ···················· | ···················· | ···················· | ···················· | ···················· | ···················· |
| E | 2168 | hCoV-19/Finland/16A24S14/2020\|EPI_ISL_481566\|2020- | ···················· | ···················· | ···················· | ···················· | ···················· | ···················· |
| E | 2169 | hCoV-19/Finland/16A25S14/2020\|EPI_ISL_481567\|2020- | ···················· | ···················· | ···················· | ···················· | ···················· | ···················· |
| E | 2170 | hCoV-19/Finland/16A68S11/2020\|EPI_ISL_481568\|2020- | ···················· | ···················· | ···················· | ···················· | ···················· | ···················· |
| E | 2171 | hCoV-19/Finland/16M18/2020\|EPI_ISL_481569\|2020-03- | ···················· | ···················· | ···················· | ···················· | ···················· | ···················· |
| E | 2172 | hCoV-19/Finland/16M32/2020\|EPI_ISL_481570\|2020-03- | ···················· | ···················· | ···················· | ···················· | ···················· | ···················· |
| E | 2173 | hCoV-19/Finland/16May68S11/2020\|EPI_ISL_481571\|202 | ···················· | ···················· | ···················· | ···················· | ···················· | ···················· |
| E | 2174 | hCoV-19/Finland/17A10S4/2020\|EPI_ISL_481572\|2020-0 | ···················· | ···················· | ···················· | ···················· | ···················· | ···················· |
| E | 2175 | hCoV-19/Finland/17A14S4/2020\|EPI_ISL_481573\|2020-0 | ···················· | ···················· | ···················· | ···················· | ···················· | ···················· |
| E | 2176 | hCoV-19/Finland/17A37S12/2020\|EPI_ISL_481574\|2020- | ···················· | ···················· | ···················· | ···················· | ···················· | ···················· |
| E | 2177 | hCoV-19/Finland/17A38S4/2020\|EPI_ISL_481575\|2020-0 | ···················· | ···················· | ···················· | ···················· | ···················· | ···················· |
| E | 2178 | hCoV-19/Finland/17A44S4/2020\|EPI_ISL_481576\|2020-0 | ···················· | ···················· | ···················· | ···················· | ···················· | ···················· |
| E | 2179 | hCoV-19/Finland/17A46S12/2020\|EPI_ISL_481577\|2020- | ···················· | ···················· | ···················· | ···················· | ···················· | ···················· |
| E | 2180 | hCoV-19/Finland/17A47S12/2020\|EPI_ISL_481578\|2020- | ···················· | ···················· | ···················· | ···················· | ···················· | ···················· |
| E | 2181 | hCoV-19/Finland/17A54S4/2020\|EPI_ISL_481579\|2020-0 | ···················· | ···················· | ···················· | ···················· | ···················· | ···················· |
| E | 2182 | hCoV-19/Finland/17A62S12/2020\|EPI_ISL_481580\|2020- | ···················· | ···················· | ···················· | ···················· | ···················· | ···················· |
| E | 2183 | hCoV-19/Finland/17A63S4/2020\|EPI_ISL_481581\|2020-0 | ···················· | ···················· | ···················· | ···················· | ···················· | ···················· |
| E | 2184 | hCoV-19/Finland/17A92S12/2020\|EPI_ISL_481593\|2020- | ···················· | ···················· | ···················· | ···················· | ···················· | ···················· |
| E | 2185 | hCoV-19/Finland/17A93S13/2020\|EPI_ISL_481594\|2020- | ···················· | ···················· | ···················· | ···················· | ···················· | ···················· |
| E | 2186 | hCoV-19/Finland/20A16S7/2020\|EPI_ISL_481595\|2020-0 | ···················· | ···················· | ···················· | ···················· | ···················· | ···················· |
| E | 2187 | hCoV-19/Finland/20A21S5/2020\|EPI_ISL_481596\|2020-0 | ···················· | ···················· | ···················· | ···················· | ···················· | ···················· |
| E | 2188 | hCoV-19/Finland/20A40S5/2020\|EPI_ISL_481597\|2020-0 | ···················· | ···················· | ···················· | ···················· | ···················· | ···················· |
| E | 2189 | hCoV-19/Finland/20A45S5/2020\|EPI_ISL_481598\|2020-0 | ···················· | ···················· | ···················· | ···················· | ···················· | ···················· |
| E | 2190 | hCoV-19/Finland/20A48S5/2020\|EPI_ISL_481599\|2020-0 | ···················· | ···················· | ···················· | ···················· | ···················· | ···················· |
| E | 2191 | hCoV-19/South Africa/KRISP-0203/2020\|EPI_ISL_48285 | ···················· | ···················· | ···················· | ···················· | ···················· | ···················· |
| E | 2192 | hCoV-19/South Africa/KRISP-0266/2020\|EPI_ISL_48285 | ···················· | ···················· | ···················· | ···················· | ···················· | ···················· |
| E | 2193 | hCoV-19/South Africa/KRISP-0275/2020\|EPI_ISL_48286 | ···················· | ···················· | ···················· | ···················· | ···················· | ···················· |
| E | 2194 | hCoV-19/Senegal/64771/2020\|EPI_ISL_482874\|2020-06- | ···················· | ···················· | ···················· | ···················· | ···················· | ···················· |
| E | 2195 | hCoV-19/Senegal/68550/2020\|EPI_ISL_482875\|2020-06- | ···················· | ···················· | ···················· | ···················· | ···················· | ···················· |
| E | 2196 | hCoV-19/Senegal/68548/2020\|EPI_ISL_482876\|2020-06- | ···················· | ···················· | ···················· | ···················· | ···················· | ···················· |
| E | 2197 | hCoV-19/Senegal/68488/2020\|EPI_ISL_482877\|2020-06- | ···················· | ···················· | ···················· | ···················· | ···················· | ···················· |
| E | 2198 | hCoV-19/Senegal/68458/2020\|EPI_ISL_482878\|2020-06- | ···················· | ···················· | ···················· | ···················· | ···················· | ···················· |
| E | 2199 | hCoV-19/France/OCC-21/2020\|EPI_ISL_482879\|2020-06- | ···················· | ···················· | ···················· | ···················· | ···················· | ···················· |
| E | 2200 | hCoV-19/France/OCC-22/2020\|EPI_ISL_482880\|2020-06- | ···················· | ···················· | ···················· | ···················· | ···················· | ···················· |
| E | 2201 | hCoV-19/France/OCC-23/2020\|EPI_ISL_482881\|2020-06- | ···················· | ···················· | ···················· | ···················· | ···················· | ···················· |
| E | 2202 | hCoV-19/France/OCC-24/2020\|EPI_ISL_482882\|2020-06- | ···················· | ···················· | ···················· | ···················· | ···················· | ···················· |
| E | 2203 | hCoV-19/France/OCC-25/2020\|EPI_ISL_482883\|2020-06- | ···················· | ···················· | ···················· | ···················· | ···················· | ···················· |
| E | 2204 | hCoV-19/France/OCC-26/2020\|EPI_ISL_482884\|2020-06- | ···················· | ···················· | ···················· | ···················· | ···················· | ···················· |
| E | 2205 | hCoV-19/France/OCC-27/2020\|EPI_ISL_482885\|2020-06- | ···················· | ···················· | ···················· | ···················· | ···················· | ···················· |
| E | 2206 | hCoV-19/France/OCC-28/2020\|EPI_ISL_482886\|2020-06- | ···················· | ···················· | ···················· | ···················· | ···················· | ···················· |
| E | 2207 | hCoV-19/France/OCC-29/2020\|EPI_ISL_482887\|2020-06- | ···················· | ···················· | ···················· | ···················· | ···················· | ···················· |
| E | 2208 | hCoV-19/France/OCC-30/2020\|EPI_ISL_482888\|2020-06- | ···················· | ···················· | ···················· | ···················· | ···················· | ···················· |
| E | 2209 | hCoV-19/France/OCC-31/2020\|EPI_ISL_482889\|2020-06- | ···················· | ···················· | ···················· | ···················· | ···················· | ···················· |
| E | 2210 | hCoV-19/England/CAMB-1B3E89/2020\|EPI_ISL_492200\|20 | ···················· | ···················· | ···················· | ···················· | ···················· | ···················· |
| E | 2211 | hCoV-19/England/CAMB-1B39B5/2020\|EPI_ISL_492201\|20 | ···················· | ···················· | ···················· | ···················· | ···················· | ···················· |
| E | 2212 | hCoV-19/England/BRIS-13282A/2020\|EPI_ISL_492203\|20 | ···················· | ···················· | ···················· | ···················· | ···················· | ···················· |
| E | 2213 | hCoV-19/England/CAMB-1B47C9/2020\|EPI_ISL_492204\|20 | ···················· | ···················· | ···················· | ···················· | ···················· | ···················· |
| E | 2214 | hCoV-19/England/CAMB-1B3D9B/2020\|EPI_ISL_492205\|20 | ···················· | ···················· | ···················· | ···················· | ···················· | ···················· |
| E | 2215 | hCoV-19/England/CAMB-1B44D1/2020\|EPI_ISL_492206\|20 | ···················· | ···················· | ···················· | ···················· | ···················· | ···················· |
| E | 2216 | hCoV-19/England/NORT-2931FE/2020\|EPI_ISL_492207\|20 | ···················· | ···················· | ···················· | ···················· | ···················· | ···················· |
| E | 2217 | hCoV-19/England/NORT-293316/2020\|EPI_ISL_492208\|20 | ···················· | ···················· | ···················· | ···················· | ···················· | ···················· |
| E | 2218 | hCoV-19/England/NORT-293361/2020\|EPI_ISL_492209\|20 | ···················· | ···················· | ···················· | ···················· | ···················· | ···················· |
| E | 2219 | hCoV-19/England/BRIS-18545D6/2020\|EPI_ISL_492211\|2 | ···················· | ···················· | ···················· | ···················· | ···················· | ···················· |
| E | 2220 | hCoV-19/England/BRIS-1853346/2020\|EPI_ISL_492213\|2 | ···················· | ···················· | ···················· | ···················· | ···················· | ···················· |
| E | 2221 | hCoV-19/England/BRIS-185316A/2020\|EPI_ISL_492214\|2 | ···················· | ···················· | ···················· | ···················· | ···················· | ···················· |
| E | 2222 | hCoV-19/England/BRIS-1853416/2020\|EPI_ISL_492215\|2 | ···················· | ···················· | ···················· | ···················· | ···················· | ···················· |
| E | 2223 | hCoV-19/England/BRIS-18539CC/2020\|EPI_ISL_492217\|2 | ···················· | ···················· | ···················· | ···················· | ···················· | ···················· |
| E | 2224 | hCoV-19/England/BRIS-1853829/2020\|EPI_ISL_492219\|2 | ···················· | ···················· | ···················· | ···················· | ···················· | ···················· |
| E | 2225 | hCoV-19/England/BRIS-185381A/2020\|EPI_ISL_492220\|2 | ···················· | ···················· | ···················· | ···················· | ···················· | ···················· |
| E | 2226 | hCoV-19/England/BRIS-1853AAB/2020\|EPI_ISL_492222\|2 | ···················· | ···················· | ···················· | ···················· | ···················· | ···················· |
| E | 2227 | hCoV-19/England/BRIS-18531E2/2020\|EPI_ISL_492223\|2 | ···················· | ···················· | ···················· | ···················· | ···················· | ···················· |
| E | 2228 | hCoV-19/England/BRIS-1853CF0/2020\|EPI_ISL_492225\|2 | ···················· | ···················· | ···················· | ···················· | ···················· | ···················· |
| E | 2229 | hCoV-19/England/BRIS-1853CD2/2020\|EPI_ISL_492226\|2 | ···················· | ···················· | ···················· | ···················· | ···················· | ···················· |
| E | 2230 | hCoV-19/England/BRIS-1854372/2020\|EPI_ISL_492229\|2 | ···················· | ···················· | ···················· | ···················· | ···················· | ···················· |
| E | 2231 | hCoV-19/England/BRIS-18540B7/2020\|EPI_ISL_492230\|2 | ···················· | ···················· | ···················· | ···················· | ···················· | ···················· |
| E | 2232 | hCoV-19/England/BRIS-1853E09/2020\|EPI_ISL_492232\|2 | ···················· | ·A·················· | ···················· | ···················· | ···················· | ···················· |
| E | 2233 | hCoV-19/England/BRIS-18540E4/2020\|EPI_ISL_492233\|2 | ···················· | ···················· | ···················· | ···················· | ···················· | ···················· |
| E | 2234 | hCoV-19/England/BRIS-1853D66/2020\|EPI_ISL_492235\|2 | ···················· | ···················· | ···················· | ···················· | ···················· | ···················· |
| E | 2235 | hCoV-19/England/BRIS-1854327/2020\|EPI_ISL_492238\|2 | ···················· | ···················· | ···················· | ···················· | ···················· | ···················· |
| E | 2236 | hCoV-19/England/BRIS-1853F15/2020\|EPI_ISL_492239\|2 | ···················· | ···················· | ···················· | ···················· | ···················· | ···················· |
| E | 2237 | hCoV-19/England/BRIS-18538FC/2020\|EPI_ISL_492240\|2 | ···················· | ···················· | ···················· | ···················· | ···················· | ···················· |

| | | | | | | | | |
|---|---|---|---|---|---|---|---|---|
| E | 2238 hCoV-19/England/BRIS-1853EFA/2020\|EPI_ISL_492252\|2 | ···················· | ···················· | ···················· | ···················· | ···················· | ···················· | |
| E | 2239 hCoV-19/England/BRIS-1854336/2020\|EPI_ISL_492253\|2 | ···················· | ···················· | ···················· | ···················· | ···················· | ···················· | |
| E | 2240 hCoV-19/England/BRIS-1853B8A/2020\|EPI_ISL_492254\|2 | ···················· | ···················· | ···················· | ···················· | ···················· | ···················· | |
| E | 2241 hCoV-19/England/BRIS-1853AD8/2020\|EPI_ISL_492256\|2 | ···················· | ····A··············· | ···················· | ···················· | ···················· | ···················· | |
| E | 2242 hCoV-19/England/BRIS-1853E54/2020\|EPI_ISL_492258\|2 | ···················· | ···················· | ···················· | ···················· | ···················· | ···················· | |
| E | 2243 hCoV-19/England/BRIS-18531F1/2020\|EPI_ISL_492261\|2 | ···················· | ···················· | ···················· | ···················· | ···················· | ···················· | |
| E | 2244 hCoV-19/England/BRIS-18538DE/2020\|EPI_ISL_492262\|2 | ···················· | ···················· | ···················· | ···················· | ···················· | ···················· | |
| E | 2245 hCoV-19/England/BRIS-1853276/2020\|EPI_ISL_492263\|2 | ···················· | ···················· | ···················· | ···················· | ···················· | ···················· | |
| E | 2246 hCoV-19/England/BRIS-185457C/2020\|EPI_ISL_492264\|2 | ···················· | ···················· | ···················· | ···················· | ···················· | ···················· | |
| E | 2247 hCoV-19/England/BRIS-185413C/2020\|EPI_ISL_492265\|2 | ···················· | ···················· | ···················· | ···················· | ···················· | ···················· | |
| E | 2248 hCoV-19/England/BRIS-1853AF6/2020\|EPI_ISL_492266\|2 | ···················· | ···················· | ···················· | ···················· | ···················· | ···················· | |
| E | 2249 hCoV-19/England/BRIS-1853768/2020\|EPI_ISL_492268\|2 | ···················· | ···················· | ···················· | ···················· | ···················· | ···················· | |
| E | 2250 hCoV-19/England/BRIS-1853E90/2020\|EPI_ISL_492270\|2 | ···················· | ···················· | ···················· | ···················· | ···················· | ···················· | |
| E | 2251 hCoV-19/England/BRIS-185412D/2020\|EPI_ISL_492271\|2 | ···················· | ···················· | ···················· | ···················· | ···················· | ···················· | |
| E | 2252 hCoV-19/England/NORT-29DEDF/2020\|EPI_ISL_493500\|20 | ···················· | ···················· | ···················· | ···················· | ···················· | ···················· | |
| E | 2253 hCoV-19/England/NORT-29DF45/2020\|EPI_ISL_493501\|20 | ···················· | ···················· | ···················· | ···················· | ···················· | ···················· | |
| E | 2254 hCoV-19/England/NORT-29DF72/2020\|EPI_ISL_493502\|20 | ···················· | ···················· | ···················· | ···················· | ···················· | ···················· | |
| E | 2255 hCoV-19/England/NORT-29E014/2020\|EPI_ISL_493504\|20 | ···················· | ···················· | ···················· | ···················· | ···················· | ···················· | |
| E | 2256 hCoV-19/England/NORT-29E023/2020\|EPI_ISL_493505\|20 | ···················· | ···················· | ···················· | ···················· | ···················· | ···················· | |
| E | 2257 hCoV-19/England/NORT-29E08D/2020\|EPI_ISL_493506\|20 | ···················· | ···················· | ···················· | ···················· | ···················· | ···················· | |
| E | 2258 hCoV-19/England/NORT-29E14E/2020\|EPI_ISL_493508\|20 | ···················· | ·A·················· | ···················· | ···················· | ···················· | ···················· | |
| E | 2259 hCoV-19/England/NORT-29E20F/2020\|EPI_ISL_493509\|20 | ···················· | ···················· | ···················· | ···················· | ···················· | ···················· | |
| E | 2260 hCoV-19/England/NORT-29E25A/2020\|EPI_ISL_493510\|20 | ···················· | ···················· | ···················· | ···················· | ···················· | ···················· | |
| E | 2261 hCoV-19/England/NORT-29E31B/2020\|EPI_ISL_493511\|20 | ···················· | ···················· | ···················· | ···················· | ···················· | ···················· | |
| E | 2262 hCoV-19/England/NORT-29E366/2020\|EPI_ISL_493512\|20 | ···················· | ···················· | ···················· | ···················· | ···················· | ···················· | |
| E | 2263 hCoV-19/England/NORT-29E3A2/2020\|EPI_ISL_493513\|20 | ···················· | ···················· | ···················· | ···················· | ···················· | ···················· | |
| E | 2264 hCoV-19/England/NORT-29E3FD/2020\|EPI_ISL_493516\|20 | ···················· | ···················· | ···················· | ···················· | ···················· | ···················· | |
| E | 2265 hCoV-19/England/NORT-29E490/2020\|EPI_ISL_493517\|20 | ···················· | ···················· | ···················· | ···················· | ···················· | ···················· | |
| E | 2266 hCoV-19/England/NORT-29E542/2020\|EPI_ISL_493518\|20 | ···················· | ···················· | ···················· | ···················· | ···················· | ···················· | |
| E | 2267 hCoV-19/England/NORT-29E5AC/2020\|EPI_ISL_493519\|20 | ········T··········· | ···················· | ···················· | ···················· | ···················· | ···················· | |
| E | 2268 hCoV-19/England/NORT-29E5D9/2020\|EPI_ISL_493521\|20 | ···················· | ···················· | ···················· | ···················· | ···················· | ···················· | |
| E | 2269 hCoV-19/England/NORT-29E612/2020\|EPI_ISL_493522\|20 | ···················· | ·A·················· | ···················· | ···················· | ···················· | ···················· | |
| E | 2270 hCoV-19/England/NORT-29E630/2020\|EPI_ISL_493523\|20 | ···················· | ···················· | ···················· | ···················· | ···················· | ···················· | |
| E | 2271 hCoV-19/England/NORT-29E6E5/2020\|EPI_ISL_493525\|20 | ···················· | ···················· | ···················· | ···················· | ···················· | ···················· | |
| E | 2272 hCoV-19/England/NORT-29E72E/2020\|EPI_ISL_493526\|20 | ···················· | ···················· | ···················· | ···················· | ···················· | ···················· | |
| E | 2273 hCoV-19/England/NORT-29E7A6/2020\|EPI_ISL_493527\|20 | ···················· | ···················· | ···················· | ···················· | ···················· | ···················· | |
| E | 2274 hCoV-19/England/NORT-29E849/2020\|EPI_ISL_493529\|20 | ···················· | ···················· | ···················· | ···················· | ···················· | ···················· | |
| E | 2275 hCoV-19/England/NORT-29E885/2020\|EPI_ISL_493530\|20 | ···················· | ·A·················· | ···················· | ···················· | ···················· | ···················· | |
| E | 2276 hCoV-19/England/NORT-29E8EF/2020\|EPI_ISL_493532\|20 | ···················· | ···················· | ···················· | ···················· | ···················· | ···················· | |
| E | 2277 hCoV-19/England/NORT-29E928/2020\|EPI_ISL_493533\|20 | ···················· | ···················A | ···················· | ···················· | ···················· | ···················· | 1 |
| E | 2278 hCoV-19/England/NORT-29E982/2020\|EPI_ISL_493534\|20 | ···················· | ···················· | ···················· | ···················· | ···················· | ···················· | |
| E | 2279 hCoV-19/England/NORT-29EBC8/2020\|EPI_ISL_493535\|20 | ···················· | ···················· | ···················· | ···················· | ···················· | ···················· | |
| E | 2280 hCoV-19/England/NORT-29ECB6/2020\|EPI_ISL_493536\|20 | ···················· | ···················· | ···················· | ···················· | ···················· | ···················· | |
| E | 2281 hCoV-19/England/NORT-29ECC5/2020\|EPI_ISL_493537\|20 | ···················· | ···················· | ···················· | ···················· | ···················· | ···················· | |
| E | 2282 hCoV-19/England/NORT-29EE29/2020\|EPI_ISL_493539\|20 | ···················· | ···················· | ···················· | ···················· | ···················· | ···················· | |
| E | 2283 hCoV-19/England/NORT-29EE56/2020\|EPI_ISL_493540\|20 | ···················· | ···················· | ···················· | ···················· | ···················· | ···················· | |
| E | 2287 hCoV-19/England/NOTT-1127D8/2020\|EPI_ISL_493548\|20 | ···················· | ···················· | ···················· | ···················· | ···················· | ···················· | |
| E | 2288 hCoV-19/England/NOTT-112811/2020\|EPI_ISL_493549\|20 | ···················· | ···················· | ···················· | ···················· | ···················· | ···················· | |
| E | 2289 hCoV-19/England/NOTT-112820/2020\|EPI_ISL_493550\|20 | ···················· | ···················· | ···················· | ···················· | ···················· | ···················· | |
| E | 2290 hCoV-19/England/NOTT-11283F/2020\|EPI_ISL_493551\|20 | ···················· | ···················· | ···················· | ···················· | ···················· | ···················· | |
| E | 2291 hCoV-19/England/NOTT-11285D/2020\|EPI_ISL_493553\|20 | ···················· | ···················· | ···················· | ···················· | ···················· | ···················· | |
| E | 2292 hCoV-19/England/NOTT-11286C/2020\|EPI_ISL_493554\|20 | ···················· | ···················· | ···················· | ···················· | ···················· | ···················· | |
| E | 2293 hCoV-19/England/NOTT-11287B/2020\|EPI_ISL_493555\|20 | ···················· | ····A··············· | ···················· | ···················· | ···················· | ···················· | |
| E | 2294 hCoV-19/England/NOTT-112899/2020\|EPI_ISL_493556\|20 | ···················· | ···················· | ···················· | ···················· | ···················· | ···················· | |
| E | 2295 hCoV-19/England/NOTT-1128D5/2020\|EPI_ISL_493558\|20 | ···················· | ···················· | ···················· | ···················· | ···················· | ···················· | |
| E | 2296 hCoV-19/England/NOTT-1128F3/2020\|EPI_ISL_493560\|20 | ···················· | ···················· | ···················· | ···················· | ···················· | ···················· | |
| E | 2297 hCoV-19/England/NOTT-11293C/2020\|EPI_ISL_493564\|20 | ···················· | ···················· | ···················· | ···················· | ···················· | ···················· | |
| E | 2298 hCoV-19/England/NOTT-112969/2020\|EPI_ISL_493567\|20 | ···················· | ···················· | ···················· | ···················· | ···················· | ···················· | |
| E | 2299 hCoV-19/England/NOTT-112978/2020\|EPI_ISL_493568\|20 | ···················· | ···················· | ···················· | ···················· | ···················· | ···················· | |
| E | 2300 hCoV-19/England/NOTT-1129B4/2020\|EPI_ISL_493570\|20 | ···················· | ···················· | ···················· | ···················· | ···················· | ···················· | |
| E | 2301 hCoV-19/England/NOTT-1129C3/2020\|EPI_ISL_493571\|20 | ···················· | ···················· | ···················· | ···················· | ···················· | ···················· | |
| E | 2302 hCoV-19/England/NOTT-112ADF/2020\|EPI_ISL_493584\|20 | ···················· | ···················· | ···················· | ···················· | ···················· | ···················· | |
| E | 2303 hCoV-19/England/NOTT-112B45/2020\|EPI_ISL_493590\|20 | ···················· | ···················· | ···················· | ···················· | ···················· | ···················· | |
| E | 2304 hCoV-19/England/NOTT-112B54/2020\|EPI_ISL_493591\|20 | ···················· | ···················· | ···················· | ···················· | ···················· | ···················· | |
| E | 2305 hCoV-19/Japan/PG-0060/2020\|EPI_ISL_479822\|2020-03- | ···················· | ···················· | ···················· | ···················· | ···················· | ···················· | |
| E | 2306 hCoV-19/Japan/PG-0063/2020\|EPI_ISL_479823\|2020-03- | ···················· | ···················· | ···················· | ···················· | ···················· | ···················· | |
| E | 2307 hCoV-19/Japan/PG-0066/2020\|EPI_ISL_479824\|2020-02- | ···················· | ···················· | ···················· | ···················· | ···················· | ···················· | |
| E | 2308 hCoV-19/Japan/PG-0068/2020\|EPI_ISL_479825\|2020-02 | ···················· | ···················· | ···················· | ···················· | ···················· | ···················· | |
| E | 2309 hCoV-19/Japan/PG-0071/2020\|EPI_ISL_479826\|2020-02- | ···················· | ···················· | ···················· | ···················· | ···················· | ···················· | |
| E | 2310 hCoV-19/Japan/PG-0072/2020\|EPI_ISL_479827\|2020-02- | ···················· | ···················· | ···················· | ···················· | ···················· | ···················· | |
| E | 2311 hCoV-19/Japan/PG-0074/2020\|EPI_ISL_479828\|2020-02- | ···················· | ···················· | ···················· | ···················· | ···················· | ···················· | |
| E | 2312 hCoV-19/Japan/PG-0076/2020\|EPI_ISL_479829\|2020-02- | ···················· | ···················· | ···················· | ···················· | ···················· | ···················· | |
| E | 2313 hCoV-19/Japan/PG-0077/2020\|EPI_ISL_479830\|2020-02- | ···················· | ···················· | ···················· | ···················· | ···················· | ···················· | |
| E | 2314 hCoV-19/Japan/PG-0080/2020\|EPI_ISL_479831\|2020-03- | ···················· | ···················· | ···················· | ···················· | ···················· | ···················· | |
| E | 2315 hCoV-19/Japan/PG-0081/2020\|EPI_ISL_479832\|2020-03- | ···················· | ···················· | ···················· | ···················· | ···················· | ···················· | |
| E | 2316 hCoV-19/Japan/PG-0082/2020\|EPI_ISL_479833\|2020-03- | ···················· | ···················· | ···················· | ···················· | ···················· | ···················· | |
| E | 2317 hCoV-19/Japan/PG-0084/2020\|EPI_ISL_479834\|2020-03- | ···················· | ···················· | ···················· | ···················· | ···················· | ···················· | |
| E | 2318 hCoV-19/Japan/PG-0087/2020\|EPI_ISL_479835\|2020-03- | ···················· | ···················· | ···················· | ···················· | ···················· | ···················· | |
| E | 2319 hCoV-19/Japan/PG-0089/2020\|EPI_ISL_479836\|2020-03- | ···················· | ···················· | ···················· | ···················· | ···················· | ···················· | |
| E | 2320 hCoV-19/Japan/PG-0091/2020\|EPI_ISL_479837\|2020-03- | ···················· | ···················· | ···················· | ···················· | ···················· | ···················· | |

| | | | | | | | | |
|---|---|---|---|---|---|---|---|---|
| E | 2321 | hCoV-19/Japan/PG-0092/2020\|EPI_ISL_479838\|2020-03- | ···················· | ···················· | ····················· | ···················· | ···················· | ···················· |
| E | 2322 | hCoV-19/Japan/PG-0095/2020\|EPI_ISL_479839\|2020-03- | ···················· | ···················· | ····················· | ···················· | ···················· | ···················· |
| E | 2323 | hCoV-19/Japan/PG-0097/2020\|EPI_ISL_479840\|2020-03- | ···················· | ···················· | ····················· | ···················· | ···················· | ···················· |
| E | 2324 | hCoV-19/Japan/PG-0098/2020\|EPI_ISL_479841\|2020-03- | ···················· | ···················· | ····················· | ···················· | ···················· | ···················· |
| E | 2325 | hCoV-19/Japan/PG-0099/2020\|EPI_ISL_479842\|2020-03- | ···················· | ···················· | ····················· | ···················· | ···················· | ···················· |
| E | 2326 | hCoV-19/Japan/PG-0100/2020\|EPI_ISL_479843\|2020-03- | ···················· | ···················· | ····················· | ···················· | ···················· | ···················· |
| E | 2327 | hCoV-19/Japan/PG-0104/2020\|EPI_ISL_479844\|2020-03- | ···················· | ···················· | ····················· | ···················· | ···················· | ···················· |
| E | 2328 | hCoV-19/Japan/PG-0106/2020\|EPI_ISL_479845\|2020-03- | ···················· | ···················· | ····················· | ···················· | ···················· | ···················· |
| E | 2329 | hCoV-19/Japan/PG-0108/2020\|EPI_ISL_479846\|2020-03- | ···················· | ···················· | ····················· | ···················· | ···················· | ···················· |
| E | 2330 | hCoV-19/Japan/PG-0110/2020\|EPI_ISL_479847\|2020-03- | ···················· | ···················· | ····················· | ···················· | ···················· | ···················· |
| E | 2331 | hCoV-19/Japan/PG-0111/2020\|EPI_ISL_479848\|2020-03- | ···················· | ···················· | ····················· | ···················· | ···················· | ···················· |
| E | 2332 | hCoV-19/Japan/PG-0113/2020\|EPI_ISL_479849\|2020-03- | ···················· | ···················· | ····················· | ···················· | ···················· | ···················· |
| E | 2333 | hCoV-19/Japan/PG-0114/2020\|EPI_ISL_479850\|2020-02- | ···················· | ···················· | ····················· | ···················· | ···················· | ···················· |
| E | 2334 | hCoV-19/Japan/PG-0115/2020\|EPI_ISL_479851\|2020-02- | ···················· | ···················· | ····················· | ···················· | ···················· | ···················· |
| E | 2335 | hCoV-19/Japan/PG-0121/2020\|EPI_ISL_479853\|2020-03- | ···················· | ···················· | ····················· | ···················· | ···················· | ···················· |
| E | 2336 | hCoV-19/Japan/PG-0122/2020\|EPI_ISL_479854\|2020-03- | ···················· | ···················· | ····················· | ···················· | ···················· | ···················· |
| E | 2337 | hCoV-19/Japan/PG-0144/2020\|EPI_ISL_479866\|2020-02 | ···················· | ···················· | ····················· | ···················· | ···················· | ···················· |
| E | 2338 | hCoV-19/Japan/PG-0145/2020\|EPI_ISL_479867\|2020-02 | ···················· | ···················· | ····················· | ···················· | ···················· | ···················· |
| E | 2339 | hCoV-19/Japan/PG-0153/2020\|EPI_ISL_479868\|2020-03- | ···················· | ···················· | ····················· | ···················· | ···················· | ···················· |
| E | 2340 | hCoV-19/Japan/PG-0158/2020\|EPI_ISL_479869\|2020-03- | ···················· | ···················· | ····················· | ···················· | ···················· | ···················· |
| E | 2341 | hCoV-19/Japan/PG-0159/2020\|EPI_ISL_479870\|2020-02- | ···················· | ·A·················· | ····················· | ···················· | ···················· | ···················· |
| E | 2342 | hCoV-19/Japan/PG-0160/2020\|EPI_ISL_479871\|2020-02- | ···················· | ·A·················· | ····················· | ···················· | ···················· | ···················· |
| E | 2343 | hCoV-19/Japan/PG-0162/2020\|EPI_ISL_479872\|2020-03- | ···················· | ···················· | ····················· | ···················· | ···················· | ···················· |
| E | 2344 | hCoV-19/Japan/PG-0163/2020\|EPI_ISL_479873\|2020-03- | ···················· | ···················· | ····················· | ···················· | ···················· | ···················· |
| E | 2345 | hCoV-19/Japan/PG-0164/2020\|EPI_ISL_479874\|2020-03- | ···················· | ···················· | ····················· | ···················· | ···················· | ···················· |
| E | 2346 | hCoV-19/Japan/PG-0165/2020\|EPI_ISL_479875\|2020-03- | ···················· | ···················· | ····················· | ···················· | ···················· | ···················· |
| E | 2347 | hCoV-19/Japan/PG-0166/2020\|EPI_ISL_479876\|2020-03- | ···················· | ···················· | ····················· | ···················· | ···················· | ···················· |
| E | 2348 | hCoV-19/Japan/PG-0167/2020\|EPI_ISL_479877\|2020-03- | ···················· | ···················· | ····················· | ···················· | ···················· | ···················· |
| E | 2349 | hCoV-19/Japan/PG-0168/2020\|EPI_ISL_479878\|2020-03- | ···················· | ···················· | ····················· | ···················· | ···················· | ···················· |
| E | 2350 | hCoV-19/Japan/PG-0170/2020\|EPI_ISL_479879\|2020-03- | ···················· | ···················· | ····················· | ···················· | ···················· | ···················· |
| E | 2351 | hCoV-19/Japan/PG-0171/2020\|EPI_ISL_479880\|2020-03- | ···················· | ···················· | ····················· | ···················· | ···················· | ···················· |
| E | 2352 | hCoV-19/Japan/PG-0172/2020\|EPI_ISL_479881\|2020-03- | ···················· | ···················· | ····················· | ···················· | ···················· | ···················· |
| E | 2353 | hCoV-19/Japan/PG-0173/2020\|EPI_ISL_479882\|2020-03- | ···················· | ···················· | ····················· | ···················· | ···················· | ···················· |
| E | 2354 | hCoV-19/Japan/PG-0174/2020\|EPI_ISL_479883\|2020-03- | ···················· | ···················· | ····················· | ···················· | ···················· | ···················· |
| E | 2355 | hCoV-19/Japan/PG-0175/2020\|EPI_ISL_479884\|2020-03- | ···················· | ···················· | ····················· | ···················· | ···················· | ···················· |
| E | 2356 | hCoV-19/Japan/PG-0176/2020\|EPI_ISL_479885\|2020-03- | ···················· | ···················· | ····················· | ···················· | ···················· | ···················· |
| E | 2357 | hCoV-19/Japan/PG-0178/2020\|EPI_ISL_479886\|2020-03- | ···················· | ···················· | ····················· | ···················· | ···················· | ···················· |
| E | 2358 | hCoV-19/Japan/PG-0179/2020\|EPI_ISL_479887\|2020-03- | ···················· | ···················· | ····················· | ···················· | ···················· | ···················· |
| E | 2359 | hCoV-19/Japan/PG-0180/2020\|EPI_ISL_479888\|2020-03- | ···················· | ···················· | ····················· | ···················· | ···················· | ···················· |
| E | 2360 | hCoV-19/Japan/PG-0181/2020\|EPI_ISL_479889\|2020-03- | ···················· | ···················· | ····················· | ···················· | ···················· | ···················· |
| E | 2361 | hCoV-19/Japan/PG-0182/2020\|EPI_ISL_479890\|2020-03- | ···················· | ···················· | ····················· | ···················· | ···················· | ···················· |
| E | 2362 | hCoV-19/Japan/PG-0185/2020\|EPI_ISL_479891\|2020-03- | ···················· | ···················· | ····················· | ···················· | ···················· | ···················· |
| E | 2363 | hCoV-19/Japan/PG-0187/2020\|EPI_ISL_479892\|2020-03- | ···················· | ···················· | ····················· | ···················· | ···················· | ···················· |
| E | 2364 | hCoV-19/Japan/PG-0188/2020\|EPI_ISL_479893\|2020-03- | ···················· | ···················· | ····················· | ···················· | ···················· | ···················· |
| E | 2365 | hCoV-19/Japan/PG-0189/2020\|EPI_ISL_479894\|2020-03- | ···················· | ···················· | ····················· | ···················· | ···················· | ···················· |
| E | 2366 | hCoV-19/Japan/PG-0190/2020\|EPI_ISL_479895\|2020-03- | ···················· | ···················· | ····················· | ···················· | ···················· | ···················· |
| E | 2367 | hCoV-19/Japan/PG-0191/2020\|EPI_ISL_479896\|2020-03- | ···················· | ···················· | ····················· | ···················· | ···················· | ···················· |
| E | 2368 | hCoV-19/Japan/PG-0192/2020\|EPI_ISL_479897\|2020-03- | ···················· | ···················· | ····················· | ···················· | ···················· | ···················· |
| E | 2369 | hCoV-19/Japan/PG-0193/2020\|EPI_ISL_479898\|2020-03- | ···················· | ···················· | ····················· | ···················· | ···················· | ···················· |
| E | 2370 | hCoV-19/Japan/PG-0194/2020\|EPI_ISL_479899\|2020-03- | ···················· | ···················· | ····················· | ···················· | ···················· | ···················· |
| E | 2371 | hCoV-19/Japan/PG-0550/2020\|EPI_ISL_480204\|2020-04- | ···················· | ···················· | ····················· | ···················· | ···················· | ···················· |
| E | 2372 | hCoV-19/Japan/PG-0552/2020\|EPI_ISL_480205\|2020-04- | ···················· | ···················· | ····················· | ···················· | ···················· | ···················· |
| E | 2373 | hCoV-19/Japan/PG-0555/2020\|EPI_ISL_480206\|2020-04- | ···················· | ···················· | ····················· | ···················· | ···················· | ···················· |
| E | 2374 | hCoV-19/Japan/PG-0556/2020\|EPI_ISL_480207\|2020-04- | ···················· | ···················· | ····················· | ···················· | ···················· | ···················· |
| E | 2375 | hCoV-19/Japan/PG-0557/2020\|EPI_ISL_480208\|2020-04- | ···················· | ···················· | ····················· | ···················· | ···················· | ···················· |
| E | 2376 | hCoV-19/Japan/PG-0559/2020\|EPI_ISL_480209\|2020-04- | ···················· | ···················· | ····················· | ···················· | ···················· | ···················· |
| E | 2377 | hCoV-19/USA/FL-BPHL-0159/2020\|EPI_ISL_480810\|2020- | ···················· | ···················· | ····················· | ···················· | ···················· | ···················· |
| E | 2378 | hCoV-19/USA/FL-BPHL-0160/2020\|EPI_ISL_480811\|2020- | ···················· | ···················· | ····················· | ···················· | ···················· | ···················· |
| E | 2379 | hCoV-19/USA/FL-BPHL-0161/2020\|EPI_ISL_480812\|2020- | ···················· | ···················· | ····················· | ···················· | ···················· | ···················· |
| E | 2380 | hCoV-19/USA/FL-BPHL-0164/2020\|EPI_ISL_480815\|2020- | ···················· | ···················· | ····················· | ···················· | ···················· | ···················· |
| E | 2381 | hCoV-19/USA/FL-BPHL-0165/2020\|EPI_ISL_480816\|2020- | ···················· | ···················· | ····················· | ···················· | ···················· | ···················· |
| E | 2382 | hCoV-19/USA/FL-BPHL-0166/2020\|EPI_ISL_480817\|2020- | ···················· | ···················· | ····················· | ···················· | ···················· | ···················· |
| E | 2383 | hCoV-19/USA/FL-BPHL-0167/2020\|EPI_ISL_480818\|2020- | ···················· | ···················· | ····················· | ···················· | ···················· | ···················· |
| E | 2385 | hCoV-19/USA/FL-BPHL-0170/2020\|EPI_ISL_480821\|2020- | ···················· | ···················· | ····················· | ···················· | ···················· | ···················· |
| E | 2386 | hCoV-19/USA/FL-BPHL-0171/2020\|EPI_ISL_480822\|2020- | ···················· | ···················· | ····················· | ···················· | ···················· | ···················· |
| E | 2387 | hCoV-19/USA/FL-BPHL-0172/2020\|EPI_ISL_480823\|2020- | ···················· | ···················· | ····················· | ···················· | ···················· | ···················· |
| E | 2388 | hCoV-19/USA/FL-BPHL-0173/2020\|EPI_ISL_480824\|2020- | ···················· | ···················· | ····················· | ···················· | ···················· | ···················· |
| E | 2389 | hCoV-19/USA/FL-BPHL-0174/2020\|EPI_ISL_480825\|2020- | ···················· | ···················· | ····················· | ···················· | ···················· | ···················· |
| E | 2390 | hCoV-19/USA/FL-BPHL-0175/2020\|EPI_ISL_480826\|2020- | ···················· | ···················· | ····················· | ···················· | ···················· | ···················· |
| E | 2391 | hCoV-19/USA/FL-BPHL-0176/2020\|EPI_ISL_480827\|2020- | ···················· | ···················· | ····················· | ···················· | ···················· | ···················· |
| E | 2392 | hCoV-19/USA/FL-BPHL-0177/2020\|EPI_ISL_480828\|2020- | ···················· | ···················· | ····················· | ···················· | ···················· | ···················· |
| E | 2393 | hCoV-19/USA/FL-BPHL-0178/2020\|EPI_ISL_480829\|2020- | ···················· | ···················· | ····················· | ···················· | ···················· | ···················· |
| E | 2394 | hCoV-19/USA/FL-BPHL-0179/2020\|EPI_ISL_480830\|2020- | ···················· | ···················· | ····················· | ···················· | ···················· | ···················· |
| E | 2395 | hCoV-19/USA/FL-BPHL-0180/2020\|EPI_ISL_480831\|2020- | ···················· | ···················· | ····················· | ···················· | ···················· | ···················· |
| E | 2396 | hCoV-19/USA/FL-BPHL-0182/2020\|EPI_ISL_480833\|2020- | ···················· | ···················· | ····················· | ···················· | ···················· | ···················· |
| E | 2397 | hCoV-19/USA/FL-BPHL-0183/2020\|EPI_ISL_480834\|2020- | ···················· | ···················· | ····················· | ···················· | ···················· | ···················· |
| E | 2398 | hCoV-19/USA/FL-BPHL-0188/2020\|EPI_ISL_480839\|2020- | ···················· | ···················· | ····················· | ···················· | ···················· | ···················· |
| E | 2399 | hCoV-19/USA/FL-BPHL-0199/2020\|EPI_ISL_480850\|2020- | ···················· | ···················· | ····················· | ···················· | ···················· | ···················· |
| E | 2400 | hCoV-19/USA/FL-BPHL-0200/2020\|EPI_ISL_480851\|2020- | ···················· | ···················· | ····················· | ···················· | ···················· | ···················· |
| E | 2401 | hCoV-19/USA/FL-BPHL-0201/2020\|EPI_ISL_480852\|2020- | ···················· | ···················· | ····················· | ···················· | ···················· | ···················· |

| | | | | | | | | |
|---|---|---|---|---|---|---|---|---|
| E | 2402 hCoV-19/USA/FL-BPHL-0204/2020\|EPI_ISL_480855\|2020- | .................... | .................... | ..................... | .................... | .................... | .................... | |
| E | 2403 hCoV-19/USA/FL-BPHL-0205/2020\|EPI_ISL_480856\|2020- | .................... | .................... | ..................... | .................... | .................... | .................... | |
| E | 2404 hCoV-19/USA/FL-BPHL-0206/2020\|EPI_ISL_480857\|2020- | .................... | .................... | ..................... | .................... | .................... | .................... | |
| E | 2405 hCoV-19/USA/FL-BPHL-0207/2020\|EPI_ISL_480858\|2020- | .................... | .................... | ..................... | .................... | .................... | .................... | |
| E | 2406 hCoV-19/USA/FL-BPHL-0208/2020\|EPI_ISL_480859\|2020- | .................... | .................... | ..................... | .................... | .................... | .................... | |
| E | 2407 hCoV-19/USA/FL-BPHL-0211/2020\|EPI_ISL_480862\|2020- | .................... | .................... | ..................... | .................... | .................... | .................... | |
| E | 2408 hCoV-19/USA/FL-BPHL-0212/2020\|EPI_ISL_480863\|2020- | .................... | .................... | ..................... | .................... | .................... | .................... | |
| E | 2409 hCoV-19/USA/FL-BPHL-0215/2020\|EPI_ISL_480866\|2020- | .................... | .................... | ..................... | .................... | .................... | .................... | |
| E | 2410 hCoV-19/USA/FL-BPHL-0219/2020\|EPI_ISL_480870\|2020- | .................... | .................... | ..................... | .................... | .................... | .................... | |
| E | 2411 hCoV-19/USA/FL-BPHL-0220/2020\|EPI_ISL_480871\|2020- | .................... | .................... | ..................... | .................... | .................... | .................... | |
| E | 2412 hCoV-19/USA/FL-BPHL-0221/2020\|EPI_ISL_480872\|2020- | .................... | .................... | ..................... | .................... | .................... | ..A................. | |
| E | 2413 hCoV-19/USA/FL-BPHL-0222/2020\|EPI_ISL_480873\|2020- | .................... | .................... | ..................... | .................... | .................... | .................... | |
| E | 2414 hCoV-19/USA/FL-BPHL-0223/2020\|EPI_ISL_480874\|2020- | .................... | .................... | ..................... | .................... | .................... | .................... | |
| E | 2415 hCoV-19/USA/FL-BPHL-0224/2020\|EPI_ISL_480875\|2020- | .................... | .................... | ..................... | .................... | .................... | .................... | |
| E | 2416 hCoV-19/USA/FL-BPHL-0225/2020\|EPI_ISL_480876\|2020- | .................... | .................... | ..................... | .................... | .................... | .................... | |
| E | 2417 hCoV-19/USA/FL-BPHL-0226/2020\|EPI_ISL_480877\|2020- | .................... | .................... | ..................... | .................... | .................... | .................... | |
| E | 2418 hCoV-19/USA/FL-BPHL-0227/2020\|EPI_ISL_480878\|2020- | .................... | .................... | ..................... | .................... | .................... | .................... | |
| E | 2419 hCoV-19/USA/FL-BPHL-0228/2020\|EPI_ISL_480879\|2020- | .................... | .................... | ..................... | .................... | .................... | .................... | |
| E | 2420 hCoV-19/USA/FL-BPHL-0229/2020\|EPI_ISL_480880\|2020- | .................... | .................... | ..................... | .................... | .................... | .................... | |
| E | 2421 hCoV-19/USA/FL-BPHL-0231/2020\|EPI_ISL_480882\|2020- | .................... | .................... | ..................... | .................... | .................... | .................... | |
| E | 2422 hCoV-19/USA/FL-BPHL-0232/2020\|EPI_ISL_480883\|2020- | .................... | .................... | ..................... | .................... | .................... | ..A................. | |
| E | 2423 hCoV-19/USA/FL-BPHL-0233/2020\|EPI_ISL_480884\|2020- | .................... | .................... | ..................... | .................... | .................... | .................... | |
| E | 2424 hCoV-19/USA/FL-BPHL-0234/2020\|EPI_ISL_480885\|2020- | .................... | .................... | ..................... | .................... | .................... | .................... | |
| E | 2425 hCoV-19/USA/FL-BPHL-0237/2020\|EPI_ISL_480888\|2020- | .................... | .................... | ..................... | .................... | .................... | .................... | |
| E | 2426 hCoV-19/USA/FL-BPHL-0238/2020\|EPI_ISL_480889\|2020- | .................... | .................... | ..................... | .................... | .................... | .................... | |
| E | 2427 hCoV-19/Finland/11A47S7/2020\|EPI_ISL_481535\|2020-0 | .................... | .................... | ..................... | .................... | .................... | .................... | |
| E | 2428 hCoV-19/Finland/11May41S7/2020\|EPI_ISL_481536\|2020 | .................... | .................... | ..................... | .................... | .................... | .................... | |
| E | 2429 hCoV-19/Finland/11May91S10/2020\|EPI_ISL_481537\|202 | .................... | .................... | ..................... | .................... | .................... | .................... | |
| E | 2430 hCoV-19/Finland/12A22SVI/2020\|EPI_ISL_481538\|2020- | .................... | .................... | ..................... | .................... | .................... | .................... | |
| E | 2431 hCoV-19/Finland/12A44SVI/2020\|EPI_ISL_481539\|2020- | .................... | .................... | ..................... | .................... | .................... | .................... | |
| E | 2432 hCoV-19/South Africa/KRISP-0280/2020\|EPI_ISL_48286 | .................... | .................... | ..................... | .................... | .................... | .................... | |
| E | 2433 hCoV-19/Australia/SAP296/2020\|EPI_ISL_483089\|2020- | .................... | .................... | ..................... | .................... | .................... | .................... | |
| E | 2434 hCoV-19/Australia/SAP303/2020\|EPI_ISL_483090\|2020- | .................... | .................... | ..................... | .................... | .................... | .................... | |
| E | 2435 hCoV-19/Australia/SAP306/2020\|EPI_ISL_483091\|2020- | .................... | .................... | ..................... | .................... | .................... | .................... | |
| E | 2436 hCoV-19/Australia/SAP309/2020\|EPI_ISL_483092\|2020- | .................... | .................... | ..................... | .................... | .................... | .................... | |
| E | 2437 hCoV-19/Australia/SAP310/2020\|EPI_ISL_483093\|2020- | .................... | .................... | ..................... | .................... | .................... | .................... | |
| E | 2438 hCoV-19/Australia/SAP312/2020\|EPI_ISL_483094\|2020- | .................... | .................... | ..................... | .................... | .................... | .................... | |
| E | 2439 hCoV-19/Australia/SAP318/2020\|EPI_ISL_483098\|2020- | .................... | .................... | ..................... | .................... | .................... | .................... | |
| E | 2440 hCoV-19/Australia/SAP319/2020\|EPI_ISL_483099\|2020- | .................... | .................... | ..................... | .................... | .................... | .................... | |
| E | 2441 hCoV-19/Northern Ireland/NIRE-1039A7/2020\|EPI_ISL_ | .................... | .................... | ..................... | .................... | .................... | .................... | |
| E | 2442 hCoV-19/Northern Ireland/NIRE-103BA1/2020\|EPI_ISL_ | .................... | .A.................. | ..................... | .................... | .................... | .................... | |
| E | 2443 hCoV-19/Northern Ireland/NIRE-103084/2020\|EPI_ISL_ | .................... | .................... | ..................... | .................... | .................... | .................... | |
| E | 2444 hCoV-19/Northern Ireland/NIRE-102AD2/2020\|EPI_ISL_ | .................... | .A.................. | ..................... | .................... | .................... | .................... | |
| E | 2445 hCoV-19/Northern Ireland/NIRE-103831/2020\|EPI_ISL_ | .................... | .................... | ..................... | .................... | .................... | .................... | |
| E | 2446 hCoV-19/Northern Ireland/NIRE-103989/2020\|EPI_ISL_ | .................... | .................... | ..................... | .................... | .................... | .................... | |
| E | 2447 hCoV-19/Northern Ireland/NIRE-103260/2020\|EPI_ISL_ | .................... | .................... | ..................... | .................... | .................... | .................... | |
| E | 2448 hCoV-19/Northern Ireland/NIRE-102D8E/2020\|EPI_ISL_ | .................... | .................... | ..................... | .................... | .................... | .................... | |
| E | 2449 hCoV-19/Northern Ireland/NIRE-103C71/2020\|EPI_ISL_ | .................... | .................... | ..................... | .................... | .................... | .................... | |
| E | 2450 hCoV-19/Northern Ireland/NIRE-103716/2020\|EPI_ISL_ | .................... | .................... | ..................... | .................... | .................... | .................... | |
| E | 2451 hCoV-19/Northern Ireland/NIRE-102C27/2020\|EPI_ISL_ | .................... | .................... | ..................... | .................... | .................... | .................... | |
| E | 2452 hCoV-19/Northern Ireland/NIRE-104575/2020\|EPI_ISL_ | .................... | .................... | ..................... | .................... | .................... | .................... | |
| E | 2453 hCoV-19/Northern Ireland/NIRE-103BCF/2020\|EPI_ISL_ | .................... | .................... | ..................... | .................... | .................... | .................... | |
| E | 2454 hCoV-19/Northern Ireland/NIRE-102C54/2020\|EPI_ISL_ | .................... | .................... | ..................... | .................... | .................... | .................... | |
| E | 2455 hCoV-19/Northern Ireland/NIRE-103B47/2020\|EPI_ISL_ | .................... | .................... | ..................... | .................... | .................... | .................... | |
| E | 2456 hCoV-19/Northern Ireland/NIRE-102DAC/2020\|EPI_ISL_ | .................... | .................... | ..................... | .................... | .................... | .................... | |
| E | 2457 hCoV-19/Northern Ireland/NIRE-103066/2020\|EPI_ISL_ | .................... | .................... | ..................... | .................... | .................... | .................... | |
| E | 2458 hCoV-19/Northern Ireland/NIRE-1001A2/2020\|EPI_ISL_ | .................... | .................... | ..................... | .................... | .................... | .................... | |
| E | 2459 hCoV-19/Northern Ireland/NIRE-103B74/2020\|EPI_ISL_ | .................... | .................... | ..................... | .................... | .................... | .................... | |
| E | 2460 hCoV-19/Northern Ireland/NIRE-1037AD/2020\|EPI_ISL_ | .................... | .................... | ..................... | .................... | .................... | .................... | |
| E | 2461 hCoV-19/Northern Ireland/NIRE-103075/2020\|EPI_ISL_ | .................... | .................... | ..................... | .................... | .................... | .................... | |
| E | 2462 hCoV-19/Spain/PV-HUB-50378813/2020\|EPI_ISL_490202\| | .................... | .................... | ..................... | .................... | .................... | .................... | |
| E | 2463 hCoV-19/Spain/PV-HUB-50378855/2020\|EPI_ISL_490203\| | .................... | .................... | ..................... | .................... | .................... | .................... | |
| E | 2464 hCoV-19/Spain/PV-HUB-50388391/2020\|EPI_ISL_490204\| | .................... | .................... | ..................... | .................... | .................... | .................... | |
| E | 2465 hCoV-19/Germany/BY-MGZ-01/2020\|EPI_ISL_490205\|2020 | .................... | .................... | ..................... | .................... | .................... | .................... | |
| E | 2466 hCoV-19/Germany/BY-MGZ-02/2020\|EPI_ISL_490206\|2020 | .................... | .................... | ..................... | .................... | .................... | .................... | |
| E | 2467 hCoV-19/Germany/BY-MGZ-03/2020\|EPI_ISL_490207\|2020 | .................... | .................... | ..................... | .................... | .................... | .................... | |
| E | 2468 hCoV-19/Germany/BY-MGZ-04/2020\|EPI_ISL_490208\|2020 | .................... | .................... | ..................... | .................... | .................... | .................... | |
| E | 2469 hCoV-19/Peru/LIM-INS-015/2020\|EPI_ISL_490209\|2020- | .................... | .................... | ..................... | .................... | .................... | .................... | |
| E | 2470 hCoV-19/Scotland/EDB1480/2020\|EPI_ISL_491500\|2020- | .................... | .A.................. | ..................... | .................... | .................... | .................... | |
| E | 2471 hCoV-19/Scotland/EDB1026/2020\|EPI_ISL_491501\|2020- | .................... | ...................A | ..................... | .................... | .................... | .................... | 1 |
| E | 2472 hCoV-19/Scotland/EDB1903/2020\|EPI_ISL_491502\|2020- | .................... | ...................A | ..................... | .................... | .................... | .................... | 1 |
| E | 2473 hCoV-19/Scotland/EDB3843/2020\|EPI_ISL_491503\|2020- | .................... | .................... | ..................... | .................... | .................... | .................... | |
| E | 2474 hCoV-19/Scotland/EDB6725/2020\|EPI_ISL_491504\|2020- | .................... | .................... | ..................... | .................... | .................... | .................... | |
| E | 2475 hCoV-19/Scotland/EDB2139/2020\|EPI_ISL_491506\|2020- | .................... | .................... | ..................... | .................... | .................... | .................... | |
| E | 2476 hCoV-19/Scotland/EDB2278/2020\|EPI_ISL_491507\|2020- | .................... | .................... | ..................... | .................... | .................... | .................... | |
| E | 2477 hCoV-19/Scotland/EDB1399/2020\|EPI_ISL_491508\|2020- | .................... | .................... | ..................... | .................... | .................... | .................... | |
| E | 2478 hCoV-19/Scotland/EDB2535/2020\|EPI_ISL_491509\|2020- | .................... | .................... | ..................... | .................... | .................... | .................... | |
| E | 2479 hCoV-19/Scotland/EDB2280/2020\|EPI_ISL_491511\|2020- | .................... | .................... | ..................... | .................... | .................... | .................... | |
| E | 2480 hCoV-19/Scotland/EDB1407/2020\|EPI_ISL_491512\|2020- | .................... | .A.................. | ..................... | .................... | .................... | .................... | |
| E | 2481 hCoV-19/Scotland/EDB2281/2020\|EPI_ISL_491513\|2020- | .................... | .................... | ..................... | .................... | .................... | .................... | |

```
E    2482 hCoV-19/Scotland/EDB2270/2020|EPI_ISL_491514|2020-   ....................  ....................  ....................  ....................  ....................  ....................
E    2483 hCoV-19/Scotland/EDB2548/2020|EPI_ISL_491515|2020-   ....................  ....................  ....................  ....................  ....................  ....................
E    2484 hCoV-19/Scotland/EDB1944/2020|EPI_ISL_491516|2020-   ....................  ....................  ....................  ....................  ....................  ....................
E    2485 hCoV-19/Scotland/EDB1915/2020|EPI_ISL_491519|2020-   ....................  .A..................  ....................  ....................  ....................  ....................
E    2486 hCoV-19/Scotland/EDB2275/2020|EPI_ISL_491520|2020-   ....................  ....................  ....................  ....................  ....................  ....................
E    2487 hCoV-19/Scotland/EDB2267/2020|EPI_ISL_491521|2020-   ....................  .A..................  ....................  ....................  ....................  ....................
E    2488 hCoV-19/Scotland/EDB1855/2020|EPI_ISL_491524|2020-   ....................  ....................  ....................  ....................  ....................  ....................
E    2489 hCoV-19/Scotland/EDB1906/2020|EPI_ISL_491525|2020-   ....................  ...................A  ....................  ....................  ....................  ....................   1
E    2490 hCoV-19/Scotland/EDB2120/2020|EPI_ISL_491527|2020-   ....................  ....................  ....................  ....................  ....................  ....................
E    2491 hCoV-19/Scotland/EDB1363/2020|EPI_ISL_491529|2020-   ....................  .A..................  ....................  ....................  ....................  ....................
E    2492 hCoV-19/Scotland/EDB1192/2020|EPI_ISL_491530|2020-   ....................  ....................  ....................  ....................  ....................  ....................
E    2493 hCoV-19/Scotland/EDB2272/2020|EPI_ISL_491531|2020-   ....................  ....................  ....................  ....................  ....................  ....................
E    2494 hCoV-19/Scotland/EDB2288/2020|EPI_ISL_491532|2020-   ....................  ....................  ....................  ....................  ....................  ....................
E    2495 hCoV-19/Scotland/EDB3721/2020|EPI_ISL_491533|2020-   ....................  ....................  ....................  ....................  ....................  ....................
E    2496 hCoV-19/Scotland/EDB2285/2020|EPI_ISL_491535|2020-   ....................  ....................  ....................  ....................  ....................  ....................
E    2497 hCoV-19/Scotland/EDB1489/2020|EPI_ISL_491539|2020-   ....................  ....................  ....................  ....................  ....................  ....................
E    2498 hCoV-19/England/BRIS-124014/2020|EPI_ISL_492800|20   ....................  ....................  ....................  ....................  ....................  ....................
E    2499 hCoV-19/England/BRIS-123CA8/2020|EPI_ISL_492801|20   ....................  ....................  ....................  ....................  ....................  ....................
E    2500 hCoV-19/England/BRIS-1242B4/2020|EPI_ISL_492803|20   ....................  .A..................  ....................  ....................  ....................  ....................
E    2501 hCoV-19/England/BRIS-1243DF/2020|EPI_ISL_492804|20   ....................  ...A................  ....................  ....................  ....................  ....................
E    2502 hCoV-19/England/BRIS-124445/2020|EPI_ISL_492806|20   ....................  ....................  ....................  ....................  ....................  ....................
E    2503 hCoV-19/England/BRIS-123F72/2020|EPI_ISL_492807|20   ....................  ....................  ....................  ....................  ....................  ....................
E    2504 hCoV-19/England/BRIS-123F90/2020|EPI_ISL_492808|20   ....................  ....................  ....................  ....................  ....................  ....................
E    2505 hCoV-19/England/BRIS-123D4B/2020|EPI_ISL_492809|20   ....................  ....................  ....................  ....................  ....................  ....................
E    2506 hCoV-19/England/BRIS-123FBE/2020|EPI_ISL_492821|20   ....................  ....................  ....................  ....................  ....................  ....................
E    2507 hCoV-19/England/BRIS-123F63/2020|EPI_ISL_492822|20   ....................  ....................  ....................  ....................  ....................  ....................
E    2508 hCoV-19/England/BRIS-123FDC/2020|EPI_ISL_492823|20   ....................  ....................  ....................  ....................  ....................  ....................
E    2509 hCoV-19/England/BRIS-123F18/2020|EPI_ISL_492825|20   ....................  ....................  ....................  ....................  ....................  ....................
E    2510 hCoV-19/England/BRIS-12468B/2020|EPI_ISL_492826|20   ....................  ....................  ....................  ....................  ....................  ....................
E    2511 hCoV-19/England/BRIS-123F54/2020|EPI_ISL_492827|20   ....................  ....................  ....................  ....................  ....................  ....................
E    2512 hCoV-19/England/BRIS-123F27/2020|EPI_ISL_492828|20   ....................  .............A......  ....................  ....................  ....................  ....................
E    2513 hCoV-19/England/BRIS-123E1B/2020|EPI_ISL_492829|20   ....................  ....................  ....................  ....................  ....................  ....................
E    2514 hCoV-19/England/BRIS-123DC3/2020|EPI_ISL_492830|20   ....................  ....................  ....................  ....................  ....................  ....................
E    2515 hCoV-19/England/BRIS-1240E7/2020|EPI_ISL_492831|20   ....................  ....................  ....................  ....................  ....................  ....................
E    2516 hCoV-19/England/BRIS-123E75/2020|EPI_ISL_492832|20   ....................  ....................  ....................  ....................  ....................  ....................
E    2517 hCoV-19/England/BRIS-1244FA/2020|EPI_ISL_492833|20   ....................  ....................  ....................  ....................  ....................  ....................
E    2518 hCoV-19/England/BRIS-12416C/2020|EPI_ISL_492834|20   ....................  ....................  ....................  ....................  ....................  ....................
E    2519 hCoV-19/England/BRIS-124111/2020|EPI_ISL_492835|20   ....................  ....................  ....................  ....................  ....................  ....................
E    2520 hCoV-19/England/WSFT-25C0E43/2020|EPI_ISL_492836|2   ....................  ....................  ....................  ....................  ....................  ....................
E    2521 hCoV-19/England/WSFT-25C0FD7/2020|EPI_ISL_492838|2   ....................  ....................  ....................  ....................  ....................  ....................
E    2522 hCoV-19/England/WSFT-25C1000/2020|EPI_ISL_492839|2   ....................  ....................  ....................  ....................  ....................  ....................
E    2523 hCoV-19/England/LOND-124C49E/2020|EPI_ISL_492840|2   ....................  ....................  ....................  ....................  ....................  ....................
E    2524 hCoV-19/England/WSFT-25C1370/2020|EPI_ISL_492841|2   ....................  ....................  ....................  ....................  ....................  ....................
E    2525 hCoV-19/England/LOND-124C3CE/2020|EPI_ISL_492843|2   ....................  ....................  ....................  ....................  ....................  ....................
E    2526 hCoV-19/England/LOND-124C337/2020|EPI_ISL_492844|2   ....................  ....................  ....................  ....................  ....................  ....................
E    2527 hCoV-19/England/LOND-124C59B/2020|EPI_ISL_492845|2   ....................  ....................  ....................  ....................  ....................  ....................
E    2528 hCoV-19/England/LOND-124C5AA/2020|EPI_ISL_492847|2   ....................  ....................  ....................  ....................  ....................  ....................
E    2529 hCoV-19/England/WSFT-25C113A/2020|EPI_ISL_492848|2   ....................  ....................  ....................  ....................  ....................  ....................
E    2530 hCoV-19/England/WSFT-25C13CB/2020|EPI_ISL_492849|2   ....................  ....................  ....................  ....................  ....................  ....................
E    2531 hCoV-19/England/LOND-124C3DD/2020|EPI_ISL_492850|2   ....................  ....................  ....................  ....................  ....................  ....................
E    2532 hCoV-19/England/LOND-124C407/2020|EPI_ISL_492852|2   ....................  ....................  ....................  ....................  ....................  ....................
E    2533 hCoV-19/England/WSFT-25C1167/2020|EPI_ISL_492853|2   ....................  ....................  ....................  ....................  ....................  ....................
E    2534 hCoV-19/England/WSFT-25C10C4/2020|EPI_ISL_492854|2   ....................  ....................  ....................  ....................  ....................  ....................
E    2535 hCoV-19/England/LOND-124C470/2020|EPI_ISL_492856|2   ....................  ....................  ....................  ....................  ....................  ....................
E    2536 hCoV-19/England/WSFT-25C0F31/2020|EPI_ISL_492857|2   ....................  ....................  ....................  ....................  ....................  ....................
E    2537 hCoV-19/England/WSFT-25C1097/2020|EPI_ISL_492860|2   ....................  ....................  ....................  ....................  ....................  ....................
E    2538 hCoV-19/England/LOND-124C5B9/2020|EPI_ISL_492864|2   ....................  ....................  ....................  ....................  ....................  ....................
E    2539 hCoV-19/England/LOND-124C522/2020|EPI_ISL_492865|2   ....................  ....................  ....................  ....................  ....................  ....................
E    2540 hCoV-19/England/WSFT-25C0D55/2020|EPI_ISL_492866|2   ....................  ....................  ....................  ....................  ....................  ....................
E    2541 hCoV-19/England/WSFT-25C111C/2020|EPI_ISL_492867|2   ....................  ....................  ....................  ....................  ....................  ....................
E    2542 hCoV-19/England/LOND-124C5C8/2020|EPI_ISL_492868|2   ....................  ....................  ....................  ....................  ....................  ....................
E    2543 hCoV-19/England/LOND-124C452/2020|EPI_ISL_492869|2   ....................  ....................  ....................  ....................  ....................  ....................
E    2544 hCoV-19/India/GJ-GBRC312b/2020|EPI_ISL_495068|2020   ....................  ....................  ....................  ....................  ....................  ....................
E    2545 hCoV-19/India/GJ-GBRC313/2020|EPI_ISL_495069|2020-   ....................  .A..................  ....................  ....................  ....................  ...A................
E    2546 hCoV-19/India/GJ-GBRC314/2020|EPI_ISL_495070|2020-   ....................  ....................  ....................  ....................  ....................  ....................
E    2547 hCoV-19/India/GJ-GBRC315a/2020|EPI_ISL_495071|2020   ....................  ....................  ....................  ....................  ....................  ....................
E    2548 hCoV-19/India/GJ-GBRC315b/2020|EPI_ISL_495072|2020   ....................  ....................  ....................  ....................  ....................  ....................
E    2549 hCoV-19/India/GJ-GBRC316a/2020|EPI_ISL_495073|2020   ....................  ....................  ....................  ....................  ....................  ....................
E    2550 hCoV-19/India/GJ-GBRC316b/2020|EPI_ISL_495074|2020   ....................  ....................  ....................  ....................  ....................  ....................
E    2551 hCoV-19/India/GJ-GBRC318a/2020|EPI_ISL_495075|2020   ....................  ....................  ....................  ....................  ....................  ....................
E    2552 hCoV-19/India/GJ-GBRC318b/2020|EPI_ISL_495076|2020   ....................  ....................  ....................  ....................  ....................  ....................
E    2553 hCoV-19/India/GJ-GBRC319/2020|EPI_ISL_495077|2020-   ....................  ....................  ....................  ....................  ....................  ....................
E    2554 hCoV-19/India/GJ-GBRC320/2020|EPI_ISL_495078|2020-   ....................  ....................  ....................  ....................  ....................  ....................
E    2555 hCoV-19/India/GJ-GBRC323a/2020|EPI_ISL_495079|2020   ....................  .A..................  ....................  ....................  ....................  ...A................
E    2556 hCoV-19/India/GJ-GBRC323b/2020|EPI_ISL_495080|2020   ....................  ....................  ....................  ....................  ....................  ...A................
E    2557 hCoV-19/England/WSFT-25C18AE/2020|EPI_ISL_495081|2   ....................  ....................  ....................  ....................  ....................  ....................
E    2558 hCoV-19/England/WSFT-25C1686/2020|EPI_ISL_495082|2   ....................  ....................  ....................  ....................  ....................  ....................
E    2559 hCoV-19/England/WSFT-25C1598/2020|EPI_ISL_495084|2   ....................  ....................  ....................  ....................  ....................  ....................
E    2560 hCoV-19/England/WSFT-25C18BD/2020|EPI_ISL_495086|2   ....................  ....................  ....................  ....................  ....................  ....................
E    2561 hCoV-19/Japan/PG-0016/2020|EPI_ISL_479800|2020-01-   ....................  ....................  ....................  ....................  ....................  ....................
```

| | | | | | | | | |
|---|---|---|---|---|---|---|---|---|
| E | 2562 | hCoV-19/Japan/PG-0022/2020\|EPI_ISL_479801\|2020-02 | .................... | .................... | ..................... | .................... | .................... | .................... |
| E | 2563 | hCoV-19/Japan/PG-0031/2020\|EPI_ISL_479802\|2020-02- | .................... | .................... | ..................... | .................... | .................... | .................... |
| E | 2564 | hCoV-19/Japan/PG-0033/2020\|EPI_ISL_479803\|2020-02- | .................... | .................... | ..................... | .................... | .................... | .................... |
| E | 2565 | hCoV-19/Japan/PG-0036/2020\|EPI_ISL_479804\|2020-02- | .................... | .................... | ..................... | .................... | .................... | .................... |
| E | 2566 | hCoV-19/Japan/PG-0038/2020\|EPI_ISL_479805\|2020-03- | .................... | .................... | ..................... | .................... | .................... | .................... |
| E | 2567 | hCoV-19/Japan/PG-0039/2020\|EPI_ISL_479806\|2020-03- | .................... | .................... | ..................... | .................... | .................... | .................... |
| E | 2568 | hCoV-19/Japan/PG-0040/2020\|EPI_ISL_479807\|2020-03- | .................... | .................... | ..................... | .................... | .................... | .................... |
| E | 2569 | hCoV-19/Japan/PG-0041/2020\|EPI_ISL_479808\|2020-03- | .................... | .................... | ..................... | .................... | .................... | .................... |
| E | 2570 | hCoV-19/Japan/PG-0045/2020\|EPI_ISL_479809\|2020-02 | .................... | .................... | ..................... | .................... | .................... | .................... |
| E | 2571 | hCoV-19/Japan/PG-0046/2020\|EPI_ISL_479810\|2020-02 | .................... | .................... | ..................... | .................... | .................... | .................... |
| E | 2572 | hCoV-19/Japan/PG-0048/2020\|EPI_ISL_479811\|2020-02 | .................... | .................... | ..................... | .................... | .................... | .................... |
| E | 2573 | hCoV-19/Japan/PG-0049/2020\|EPI_ISL_479812\|2020-03- | .................... | .................... | ..................... | .................... | .................... | .................... |
| E | 2574 | hCoV-19/Japan/PG-0050/2020\|EPI_ISL_479813\|2020-03- | .................... | .................... | ..................... | .................... | .................... | .................... |
| E | 2575 | hCoV-19/Japan/PG-0051/2020\|EPI_ISL_479814\|2020-03- | .................... | .................... | ..................... | .................... | .................... | .................... |
| E | 2576 | hCoV-19/Japan/PG-0052/2020\|EPI_ISL_479815\|2020-03- | .................... | .................... | ..................... | .................... | .................... | .................... |
| E | 2577 | hCoV-19/Japan/PG-0053/2020\|EPI_ISL_479816\|2020-03- | .................... | .................... | ..................... | .................... | .................... | .................... |
| E | 2578 | hCoV-19/Japan/PG-0054/2020\|EPI_ISL_479817\|2020-03- | .................... | .................... | ..................... | .................... | .................... | .................... |
| E | 2579 | hCoV-19/Japan/PG-0055/2020\|EPI_ISL_479818\|2020-03- | .................... | .................... | ..................... | .................... | .................... | .................... |
| E | 2580 | hCoV-19/Japan/PG-0056/2020\|EPI_ISL_479819\|2020-03- | .................... | .................... | ..................... | .................... | .................... | .................... |
| E | 2581 | hCoV-19/Japan/PG-0057/2020\|EPI_ISL_479820\|2020-03- | .................... | .................... | ..................... | .................... | .................... | .................... |
| E | 2582 | hCoV-19/Japan/PG-0058/2020\|EPI_ISL_479821\|2020-03- | .................... | .................... | ..................... | .................... | .................... | .................... |
| E | 2583 | hCoV-19/Japan/PG-0128/2020\|EPI_ISL_479855\|2020-03- | .................... | .................... | ..................... | .................... | .................... | .................... |
| E | 2584 | hCoV-19/Japan/PG-0130/2020\|EPI_ISL_479856\|2020-03- | .................... | .................... | ..................... | .................... | .................... | .................... |
| E | 2585 | hCoV-19/Japan/PG-0131/2020\|EPI_ISL_479857\|2020-03- | .................... | .................... | ..................... | .................... | .................... | .................... |
| E | 2586 | hCoV-19/Japan/PG-0132/2020\|EPI_ISL_479858\|2020-03- | .................... | .................... | ..................... | .................... | .................... | .................... |
| E | 2587 | hCoV-19/Japan/PG-0133/2020\|EPI_ISL_479859\|2020-03- | .................... | .................... | ..................... | .................... | .................... | .................... |
| E | 2588 | hCoV-19/Japan/PG-0136/2020\|EPI_ISL_479860\|2020-03- | .................... | .................... | ..................... | .................... | .................... | .................... |
| E | 2589 | hCoV-19/Japan/PG-0138/2020\|EPI_ISL_479861\|2020-03- | .................... | .................... | ..................... | .................... | .................... | .................... |
| E | 2590 | hCoV-19/Japan/PG-0139/2020\|EPI_ISL_479862\|2020-03- | .................... | .................... | ..................... | .................... | .................... | .................... |
| E | 2591 | hCoV-19/Japan/PG-0140/2020\|EPI_ISL_479863\|2020-02- | .................... | .................... | ..................... | .................... | .................... | .................... |
| E | 2592 | hCoV-19/Japan/PG-0141/2020\|EPI_ISL_479864\|2020-02- | .................... | .................... | ..................... | .................... | .................... | .................... |
| E | 2593 | hCoV-19/Japan/PG-0143/2020\|EPI_ISL_479865\|2020-02 | .................... | .................... | ..................... | .................... | .................... | .................... |
| E | 2594 | hCoV-19/USA/FL-BPHL-0150/2020\|EPI_ISL_480801\|2020- | .................... | .................... | ..................... | .................... | .................... | .................... |
| E | 2598 | hCoV-19/USA/FL-BPHL-0155/2020\|EPI_ISL_480806\|2020- | .................... | .................... | ..................... | .................... | .................... | .................... |
| E | 2599 | hCoV-19/USA/FL-BPHL-0157/2020\|EPI_ISL_480808\|2020- | .................... | .................... | ..................... | .................... | .................... | .................... |
| E | 2601 | hCoV-19/USA/FL-BPHL-0181/2020\|EPI_ISL_480832\|2020- | .................... | .................... | ..................... | .................... | .................... | .................... |
| E | 2603 | hCoV-19/USA/FL-BPHL-0185/2020\|EPI_ISL_480836\|2020- | .................... | .................... | ..................... | .................... | .................... | .................... |
| E | 2604 | hCoV-19/USA/FL-BPHL-0186/2020\|EPI_ISL_480837\|2020- | .................... | .................... | ..................... | .................... | .................... | .................... |
| E | 2605 | hCoV-19/USA/FL-BPHL-0187/2020\|EPI_ISL_480838\|2020- | .................... | .................... | ..................... | .................... | .................... | .................... |
| E | 2606 | hCoV-19/USA/FL-BPHL-0189/2020\|EPI_ISL_480840\|2020- | .................... | .................... | ..................... | .................... | .................... | .................... |
| E | 2607 | hCoV-19/USA/FL-BPHL-0190/2020\|EPI_ISL_480841\|2020- | .................... | .................... | ..................... | .................... | .................... | .................... |
| E | 2608 | hCoV-19/USA/FL-BPHL-0191/2020\|EPI_ISL_480842\|2020- | .................... | .................... | ..................... | .................... | .................... | .................... |
| E | 2609 | hCoV-19/USA/FL-BPHL-0193/2020\|EPI_ISL_480844\|2020- | .................... | .................... | ..................... | .................... | .................... | .................... |
| E | 2610 | hCoV-19/USA/FL-BPHL-0194/2020\|EPI_ISL_480845\|2020- | .................... | .................... | ..................... | .................... | .................... | .................... |
| E | 2612 | hCoV-19/USA/FL-BPHL-0196/2020\|EPI_ISL_480847\|2020- | .................... | .................... | ..................... | .................... | .................... | .................... |
| E | 2613 | hCoV-19/USA/MN-MDH-1279/2020\|EPI_ISL_483000\|2020-0 | .................... | .................... | ..................... | .................... | .................... | .................... |
| E | 2614 | hCoV-19/USA/MN-MDH-1280/2020\|EPI_ISL_483001\|2020-0 | .................... | .................... | ..................... | .................... | .................... | .................... |
| E | 2615 | hCoV-19/USA/MN-MDH-1281/2020\|EPI_ISL_483002\|2020-0 | .................... | .................... | ..................... | .................... | .................... | .................... |
| E | 2616 | hCoV-19/USA/MN-MDH-1282/2020\|EPI_ISL_483003\|2020-0 | .................... | .................... | ..................... | .................... | .................... | .................... |
| E | 2617 | hCoV-19/USA/MN-MDH-1283/2020\|EPI_ISL_483004\|2020-0 | .................... | .................... | ..................... | .................... | .................... | .................... |
| E | 2618 | hCoV-19/USA/MN-MDH-1284/2020\|EPI_ISL_483005\|2020-0 | .................... | .................... | ..................... | .................... | .................... | .................... |
| E | 2619 | hCoV-19/USA/MN-MDH-1285/2020\|EPI_ISL_483006\|2020-0 | .................... | .................... | ..................... | .................... | .................... | .................... |
| E | 2620 | hCoV-19/USA/MN-MDH-1286/2020\|EPI_ISL_483007\|2020-0 | .................... | .................... | ..................... | .................... | .................... | .................... |
| E | 2621 | hCoV-19/USA/MN-MDH-1287/2020\|EPI_ISL_483008\|2020-0 | .................... | .................... | ..................... | .................... | .................... | .................... |
| E | 2622 | hCoV-19/USA/MN-MDH-1288/2020\|EPI_ISL_483009\|2020-0 | .................... | .................... | ..................... | .................... | .................... | .................... |
| E | 2623 | hCoV-19/USA/MN-MDH-1290/2020\|EPI_ISL_483010\|2020-0 | .................... | .................... | ..................... | .................... | .................... | .................... |
| E | 2624 | hCoV-19/USA/MN-MDH-1291/2020\|EPI_ISL_483011\|2020-0 | .................... | .................... | ..................... | .................... | .................... | .................... |
| E | 2625 | hCoV-19/USA/MN-MDH-1292/2020\|EPI_ISL_483012\|2020-0 | .................... | .................... | ..................... | .................... | .................... | .................... |
| E | 2626 | hCoV-19/USA/MN-MDH-1293/2020\|EPI_ISL_483013\|2020-0 | .................... | .................... | ..................... | .................... | .................... | .................... |
| E | 2627 | hCoV-19/USA/MN-MDH-1294/2020\|EPI_ISL_483014\|2020-0 | .................... | .................... | ..................... | .................... | .................... | .................... |
| E | 2628 | hCoV-19/USA/MN-MDH-1295/2020\|EPI_ISL_483015\|2020-0 | .................... | .................... | ..................... | .................... | .................... | .................... |
| E | 2629 | hCoV-19/USA/MN-MDH-1296/2020\|EPI_ISL_483016\|2020-0 | .................... | .................... | ..................... | .................... | .................... | .................... |
| E | 2630 | hCoV-19/USA/MN-MDH-1297/2020\|EPI_ISL_483017\|2020-0 | .................... | .................... | ..................... | .................... | .................... | .................... |
| E | 2631 | hCoV-19/USA/UT-03887/2020\|EPI_ISL_483018\|2020-05-1 | .................... | .................... | ..................... | .................... | .................... | .................... |
| E | 2632 | hCoV-19/USA/UT-04056/2020\|EPI_ISL_483024\|2020-05-1 | .................... | .................... | ..................... | .................... | .................... | .................... |
| E | 2633 | hCoV-19/USA/UT-05810/2020\|EPI_ISL_483031\|2020-05-1 | .................... | .................... | ..................... | .................... | .................... | .................... |
| E | 2634 | hCoV-19/Egypt/MASRI-009/2020\|EPI_ISL_483035\|2020-0 | .................... | .................... | ..................... | .................... | .................... | .................... |
| E | 2635 | hCoV-19/Egypt/MASRI-018/2020\|EPI_ISL_483036\|2020-0 | .................... | .................... | ..................... | .................... | .................... | .................... |
| E | 2636 | hCoV-19/Egypt/MASRI-028/2020\|EPI_ISL_483038\|2020-0 | .................... | .................... | ..................... | .................... | .................... | .................... |
| E | 2637 | hCoV-19/Spain/CT-IrsiCaixa-Own160320/2020\|EPI_ISL_ | .................... | .................... | ..................... | .................... | .................... | .................... |
| E | 2638 | hCoV-19/New Zealand/20CHL869065O/2020\|EPI_ISL_4830 | .................... | .................... | ..................... | .................... | .................... | .................... |
| E | 2639 | hCoV-19/cat/France/Env-Ba/2020\|EPI_ISL_483063\|2020 | .................... | .................... | ..................... | .................... | .................... | .................... |
| E | 2640 | hCoV-19/cat/France/Env-Di/2020\|EPI_ISL_483064\|2020 | .................... | .................... | ..................... | .................... | .................... | .................... |
| E | 2641 | hCoV-19/Brazil/RJ-DCVN01/2020\|EPI_ISL_483065\|2020- | .................... | .................... | ..................... | .................... | .................... | .................... |
| E | 2642 | hCoV-19/Australia/SAP098/2020\|EPI_ISL_483066\|2020- | .................... | .................... | ..................... | .................... | .................... | .................... |
| E | 2643 | hCoV-19/Australia/SAP195/2020\|EPI_ISL_483068\|2020- | .................... | .................... | ..................... | .................... | .................... | .................... |
| E | 2644 | hCoV-19/Australia/SAP202/2020\|EPI_ISL_483071\|2020- | .................... | .................... | ..................... | .................... | .................... | .................... |
| E | 2645 | hCoV-19/Australia/SAP234/2020\|EPI_ISL_483078\|2020- | .................... | .................... | ..................... | .................... | .................... | .................... |
| E | 2646 | hCoV-19/Australia/SAP239/2020\|EPI_ISL_483080\|2020- | .................... | .................... | ..................... | .................... | .................... | .................... |
| E | 2647 | hCoV-19/Australia/SAP254/2020\|EPI_ISL_483082\|2020- | .................... | .................... | ..................... | .................... | .................... | .................... |

```
E    2648 hCoV-19/Australia/SAP279/2020|EPI_ISL_483086|2020-   ····················  ····················  ·····················  ····················  ····················  ····················
E    2649 hCoV-19/Australia/SAP291/2020|EPI_ISL_483088|2020-   ····················  ····················  ·····················  ····················  ····················  ····················
E    2650 hCoV-19/England/NORT-29D6B9/2020|EPI_ISL_484300|20   ····················  ····················  ·····················  ····················  ····················  ····················
E    2651 hCoV-19/England/NORT-29D798/2020|EPI_ISL_484302|20   ····················  ····················  ·····················  ····················  ····················  ····················
E    2652 hCoV-19/England/NORT-29D877/2020|EPI_ISL_484304|20   ····················  ····················  ·····················  ····················  ····················  ····················
E    2653 hCoV-19/England/NORT-29D8B3/2020|EPI_ISL_484305|20   ····················  ····················  ·····················  ····················  ····················  ····················
E    2654 hCoV-19/England/NORT-29DA62/2020|EPI_ISL_484306|20   ····················  ····················  ·····················  ····················  ····················  ····················
E    2655 hCoV-19/England/NORT-29DB32/2020|EPI_ISL_484307|20   ····················  ····················  ·····················  ····················  ····················  ····················
E    2656 hCoV-19/England/NORT-29DB8D/2020|EPI_ISL_484309|20   ····················  ····················  ·····················  ····················  ····················  ····················
E    2657 hCoV-19/England/NORT-29DBAB/2020|EPI_ISL_484310|20   ····················  ····················  ·····················  ····················  ····················  ····················
E    2658 hCoV-19/England/NORT-29DC4E/2020|EPI_ISL_484312|20   ····················  ····················  ·····················  ····················  ····················  ····················
E    2659 hCoV-19/England/NORT-29DC8A/2020|EPI_ISL_484313|20   ····················  ····················  ·····················  ····················  ····················  ····················
E    2660 hCoV-19/England/NORT-29DD3C/2020|EPI_ISL_484315|20   ····················  ····················  ·····················  ····················  ····················  ····················
E    2661 hCoV-19/England/NORT-29DD4B/2020|EPI_ISL_484316|20   ····················  ····················  ·····················  ····················  ····················  ····················
E    2662 hCoV-19/England/NORT-29DDB4/2020|EPI_ISL_484317|20   ····················  ····················  ·····················  ····················  ····················  ····················
E    2663 hCoV-19/England/NORT-29DE0C/2020|EPI_ISL_484318|20   ····················  ····················  ·····················  ····················  ····················  ····················
E    2664 hCoV-19/England/NORT-29EFCC/2020|EPI_ISL_484320|20   ····················  ····················  ·····················  ····················  ····················  ····················
E    2665 hCoV-19/England/NORT-29F0AA/2020|EPI_ISL_484321|20   ····················  ····················  ·····················  ····················  ····················  ····················
E    2666 hCoV-19/England/NORT-29F0E6/2020|EPI_ISL_484322|20   ····················  ···················A  ·····················  ····················  ····················  ····················        1
E    2667 hCoV-19/England/NORT-29F1C5/2020|EPI_ISL_484323|20   ····················  ····················  ·····················  ····················  ····················  ····················
E    2668 hCoV-19/England/NORT-29F1D4/2020|EPI_ISL_484324|20   ····················  ····················  ·····················  ····················  ····················  ····················
E    2669 hCoV-19/England/NORT-29F541/2020|EPI_ISL_484325|20   ····················  ····················  ·····················  ····················  ····················  ····················
E    2670 hCoV-19/England/NORT-29F699/2020|EPI_ISL_484326|20   ····················  ·A··················  ·····················  ····················  ····················  ····················
E    2671 hCoV-19/England/NORT-29F6C6/2020|EPI_ISL_484327|20   ····················  ·A··················  ·····················  ····················  ····················  ····················
E    2672 hCoV-19/England/NOTT-10DE9F/2020|EPI_ISL_484339|20   ····················  ····················  ·····················  ····················  ····················  ····················
E    2673 hCoV-19/England/NOTT-1120FB/2020|EPI_ISL_484341|20   ····················  ····················  ·····················  ····················  ····················  ····················
E    2674 hCoV-19/England/NOTT-112116/2020|EPI_ISL_484343|20   ····················  ····················  ·····················  ····················  ····················  ····················
E    2675 hCoV-19/England/NOTT-112134/2020|EPI_ISL_484345|20   ····················  ····················  ·····················  ····················  ····················  ····················
E    2676 hCoV-19/England/NOTT-112143/2020|EPI_ISL_484346|20   ····················  ····················  ·····················  ····················  ····················  ····················
E    2677 hCoV-19/England/NOTT-112161/2020|EPI_ISL_484348|20   ····················  ····················  ·····················  ····················  ····················  ····················
E    2678 hCoV-19/England/NOTT-112170/2020|EPI_ISL_484349|20   ····················  ····················  ·····················  ····················  ····················  ····················
E    2679 hCoV-19/England/NOTT-1121CB/2020|EPI_ISL_484350|20   ····················  ····················  ·····················  ····················  ····················  ····················
E    2680 hCoV-19/England/NOTT-112204/2020|EPI_ISL_484354|20   ····················  ····················  ·····················  ····················  ····················  ····················
E    2681 hCoV-19/England/NOTT-112213/2020|EPI_ISL_484355|20   ····················  ····················  ·····················  ····················  ····················  ····················
E    2682 hCoV-19/England/NOTT-112231/2020|EPI_ISL_484357|20   ····················  ····················  ·····················  ····················  ····················  ····················
E    2683 hCoV-19/England/NOTT-112240/2020|EPI_ISL_484358|20   ····················  ····················  ·····················  ····················  ····················  ····················
E    2684 hCoV-19/England/NOTT-11226E/2020|EPI_ISL_484360|20   ····················  ····················  ·····················  ····················  ····················  ····················
E    2685 hCoV-19/England/NOTT-11227D/2020|EPI_ISL_484361|20   ····················  ····················  ·····················  ····················  ····················  ····················
E    2686 hCoV-19/England/NOTT-11228C/2020|EPI_ISL_484362|20   ····················  ····················  ·····················  ····················  ····················  ····················
E    2687 hCoV-19/England/NOTT-11229B/2020|EPI_ISL_484363|20   ····················  ····················  ·····················  ····················  ····················  ····················
E    2688 hCoV-19/England/NOTT-1122D7/2020|EPI_ISL_484366|20   ····················  ····················  ·····················  ····················  ····················  ····················
E    2689 hCoV-19/England/NOTT-1122F5/2020|EPI_ISL_484368|20   ····················  ····················  ·····················  ····················  ····················  ····················
E    2690 hCoV-19/England/NOTT-112301/2020|EPI_ISL_484369|20   ····················  ····················  ·····················  ····················  ····················  ····················
E    2691 hCoV-19/England/NOTT-112310/2020|EPI_ISL_484370|20   ····················  ····················  ·····················  ····················  ····················  ····················
E    2692 hCoV-19/England/NOTT-11232F/2020|EPI_ISL_484371|20   ····················  ····················  ·····················  ····················  ····················  ····················
E    2693 hCoV-19/England/NOTT-11233E/2020|EPI_ISL_484372|20   ····················  ····················  ·····················  ····················  ····················  ····················
E    2694 hCoV-19/England/NOTT-11235C/2020|EPI_ISL_484374|20   ····················  ····················  ·····················  ····················  ····················  ····················
E    2695 hCoV-19/England/NOTT-11237A/2020|EPI_ISL_484376|20   ····················  ····················  ·····················  ····················  ····················  ····················
E    2696 hCoV-19/England/NOTT-1123E3/2020|EPI_ISL_484377|20   ····················  ····················  ·····················  ····················  ····················  ····················
E    2697 hCoV-19/England/NOTT-1123F2/2020|EPI_ISL_484378|20   ····················  ····················  ·····················  ····················  ····················  ····················
E    2698 hCoV-19/England/NOTT-11240E/2020|EPI_ISL_484379|20   ····················  ····················  ·····················  ····················  ····················  ····················
E    2699 hCoV-19/England/NOTT-112459/2020|EPI_ISL_484381|20   ····················  ····················  ·····················  ····················  ····················  ····················
E    2700 hCoV-19/England/NOTT-112486/2020|EPI_ISL_484382|20   ····················  ····················  ·····················  ····················  ····················  ····················
E    2701 hCoV-19/England/NOTT-1124A4/2020|EPI_ISL_484384|20   ····················  ····················  ·····················  ····················  ····················  ····················
E    2702 hCoV-19/England/NOTT-11250B/2020|EPI_ISL_484388|20   ····················  ····················  ·····················  ····················  ····················  ····················
E    2703 hCoV-19/England/NOTT-11251A/2020|EPI_ISL_484389|20   ····················  ····················  ·····················  ····················  ····················  ····················
E    2704 hCoV-19/England/NOTT-112547/2020|EPI_ISL_484392|20   ····················  ····················  ·····················  ····················  ····················  ····················
E    2705 hCoV-19/England/NOTT-112556/2020|EPI_ISL_484393|20   ····················  ····················  ·····················  ····················  ····················  ····················
E    2706 hCoV-19/England/NOTT-112565/2020|EPI_ISL_484394|20   ····················  ····················  ·····················  ····················  ····················  ····················
E    2707 hCoV-19/England/NOTT-112583/2020|EPI_ISL_484396|20   ····················  ····················  ·····················  ····················  ····················  ····················
E    2708 hCoV-19/England/NOTT-1125B0/2020|EPI_ISL_484399|20   ····················  ····················  ·····················  ····················  ····················  ····················
E    2709 hCoV-19/England/200920118/2020|EPI_ISL_485604|2020   ····················  ····················  ·····················  ····················  ····················  ····················
E    2710 hCoV-19/England/202561784/2020|EPI_ISL_485605|2020   ····················  ····················  ·····················  ····················  ····················  ····················
E    2711 hCoV-19/England/202561785/2020|EPI_ISL_485606|2020   ····················  ····················  ·····················  ····················  ····················  ····················
E    2712 hCoV-19/England/202561786/2020|EPI_ISL_485607|2020   ····················  ····················  ·····················  ····················  ····················  ····················
E    2713 hCoV-19/England/202561787/2020|EPI_ISL_485608|2020   ····················  ····················  ·····················  ····················  ····················  ····················
E    2714 hCoV-19/England/202561788/2020|EPI_ISL_485609|2020   ····················  ····················  ·····················  ····················  ····················  ····················
E    2715 hCoV-19/Senegal/68349/2020|EPI_ISL_485635|2020-06-   ····················  ····················  ·····················  ····················  ····················  ····················
E    2716 hCoV-19/Japan/TK-Y17904/2020|EPI_ISL_486910|2020-0   ····················  ····················  ·····················  ····················  ····················  ····················
E    2717 hCoV-19/Japan/TK-Y17926/2020|EPI_ISL_486911|2020-0   ····················  ····················  ·····················  ····················  ····················  ····················
E    2718 hCoV-19/USA/MD-MDH-0018/2020|EPI_ISL_486912|2020-0   ····················  ····················  ·····················  ····················  ····················  ····················
E    2719 hCoV-19/USA/MD-MDH-0022/2020|EPI_ISL_486913|2020-0   ····················  ····················  ·····················  ····················  ······T·············  ····················
E    2720 hCoV-19/USA/MD-MDH-0024/2020|EPI_ISL_486914|2020-0   ····················  ····················  ·····················  ····················  ····················  ····················
E    2721 hCoV-19/USA/MD-MDH-0029/2020|EPI_ISL_486915|2020-0   ····················  ····················  ·····················  ····················  ····················  ····················
E    2722 hCoV-19/USA/MD-MDH-0037/2020|EPI_ISL_486916|2020-0   ····················  ····················  ·····················  ····················  ····················  ····················
E    2723 hCoV-19/USA/MD-MDH-0038/2020|EPI_ISL_486917|2020-0   ····················  ····················  ·····················  ····················  ····················  ····················
E    2724 hCoV-19/England/NORT-291EDB/2020|EPI_ISL_489150|20   ····················  ····················  ·····················  ····················  ····················  ····················
E    2725 hCoV-19/England/NORT-292A30/2020|EPI_ISL_489151|20   ····················  ····················  ·····················  ····················  ····················  ····················
E    2726 hCoV-19/England/NORT-291D74/2020|EPI_ISL_489153|20   ····················  ····················  ·····················  ····················  ····················  ····················
E    2727 hCoV-19/England/NORT-292511/2020|EPI_ISL_489154|20   ····················  ····················  ·····················  ····················  ····················  ····················
```

```
E    2728 hCoV-19/England/NORT-2925F3/2020|EPI_ISL_489155|20    ····················  ····················  ····················  ····················  ····················  ····················
E    2729 hCoV-19/Northern Ireland/NIRE-FEEDE/2020|EPI_ISL_4    ····················  ····················  ····················  ····················  ····················  ····················
E    2730 hCoV-19/Northern Ireland/NIRE-FCF82/2020|EPI_ISL_4    ····················  ····················  ····················  ····················  ····················  ····················
E    2731 hCoV-19/Northern Ireland/NIRE-103752/2020|EPI_ISL_    ····················  ····················  ····················  ····················  ····················  ····················
E    2732 hCoV-19/Northern Ireland/NIRE-103CCC/2020|EPI_ISL_    ····················  ····················  ····················  ····················  ····················  ····················
E    2733 hCoV-19/Northern Ireland/NIRE-102C63/2020|EPI_ISL_    ····················  ····················  ····················  ····················  ····················  ····················
E    2734 hCoV-19/Northern Ireland/NIRE-100272/2020|EPI_ISL_    ····················  ····················  ····················  ····················  ····················  ····················
E    2735 hCoV-19/Northern Ireland/NIRE-103B29/2020|EPI_ISL_    ····················  ····················  ····················  ····················  ····················  ····················
E    2736 hCoV-19/Northern Ireland/NIRE-103CBD/2020|EPI_ISL_    ····················  ····················  ····················  ····················  ····················  ····················
E    2737 hCoV-19/Northern Ireland/NIRE-103BDE/2020|EPI_ISL_    ····················  ····················  ····················  ····················  ····················  ····················
E    2738 hCoV-19/Northern Ireland/NIRE-102CAF/2020|EPI_ISL_    ····················  ····················  ····················  ····················  ····················  ····················
E    2739 hCoV-19/Northern Ireland/NIRE-FE0C9/2020|EPI_ISL_4    ····················  ····················  ····················  ····················  ····················  ····················
E    2740 hCoV-19/Bangladesh/BCSIR-NILMRC-251/2020|EPI_ISL_4    ····················  ····················  ····················  ····················  ····················  ····················
E    2741 hCoV-19/Peru/LAM-INS-035/2020|EPI_ISL_491460|2020-    ····················  ····················  ····················  ····················  ····················  ····················
E    2742 hCoV-19/Peru/LAM-INS-038/2020|EPI_ISL_491461|2020-    ····················  ····················  ····················  ····················  ····················  ····················
E    2743 hCoV-19/Peru/ICA-INS-040/2020|EPI_ISL_491462|2020-    ····················  ····················  ····················  ····················  ····················  ····················
E    2745 hCoV-19/USA/COM0001/2020|EPI_ISL_491481|2020-04-03    ····················  ·········A··········  ····················  ····················  ····················  ····················
E    2746 hCoV-19/England/BRIS-124199/2020|EPI_ISL_492791|20    ····················  ·A··················  ····················  ····················  ····················  ····················
E    2747 hCoV-19/England/BRIS-1244EB/2020|EPI_ISL_492792|20    ····················  ····················  ····················  ····················  ····················  ····················
E    2748 hCoV-19/England/BRIS-123F45/2020|EPI_ISL_492810|20    ····················  ····················  ····················  ····················  ····················  ····················
E    2749 hCoV-19/England/BRIS-1244CD/2020|EPI_ISL_492811|20    ····················  ····················  ····················  ····················  ····················  ····················
E    2750 hCoV-19/England/BRIS-12432A/2020|EPI_ISL_492814|20    ····················  ····················  ····················  ····················  ····················  ····················
E    2751 hCoV-19/England/BRIS-12414E/2020|EPI_ISL_492815|20    ····················  ····················  ····················  ····················  ····················  ····················
E    2752 hCoV-19/England/BRIS-123FFA/2020|EPI_ISL_492816|20    ····················  ····················  ····················  ····················  ····················  ····················
E    2753 hCoV-19/England/BRIS-1240AB/2020|EPI_ISL_492817|20    ····················  ····················  ····················  ····················  ····················  ····················
E    2754 hCoV-19/England/BRIS-12469A/2020|EPI_ISL_492818|20    ····················  ·············A······  ····················  ····················  ····················  ····················
E    2755 hCoV-19/England/BRIS-123CB7/2020|EPI_ISL_492819|20    ····················  ····················  ····················  ····················  ····················  ····················
E    2756 hCoV-19/Canada/LTRI-23/2020|EPI_ISL_495012|2020-03    ····················  ····················  ····················  ····················  ····················  ····················
E    2757 hCoV-19/Canada/LTRI-25/2020|EPI_ISL_495013|2020-03    ····················  ····················  ····················  ····················  ····················  ····················
E    2758 hCoV-19/India/GJ-GBRC279/2020|EPI_ISL_495016|2020-    ····················  ····················  ····················  ····················  ····················  ····················
E    2759 hCoV-19/India/GJ-GBRC280/2020|EPI_ISL_495017|2020-    ····················  ·A··················  ····················  ····················  ····················  ···A················
E    2760 hCoV-19/India/GJ-GBRC281/2020|EPI_ISL_495018|2020-    ····················  ····················  ····················  ····················  ····················  ····················
E    2761 hCoV-19/India/GJ-GBRC282/2020|EPI_ISL_495019|2020-    ····················  ·A··················  ····················  ····················  ····················  ···A················
E    2762 hCoV-19/India/GJ-GBRC283/2020|EPI_ISL_495020|2020-    ····················  ····················  ····················  ····················  ····················  ····················
E    2763 hCoV-19/India/GJ-GBRC284a/2020|EPI_ISL_495021|2020    ····················  ·A··················  ····················  ····················  ····················  ···A················
E    2764 hCoV-19/India/GJ-GBRC284b/2020|EPI_ISL_495022|2020    ····················  ·A··················  ····················  ····················  ····················  ···A················
E    2765 hCoV-19/India/GJ-GBRC285/2020|EPI_ISL_495023|2020-    ····················  ····················  ····················  ····················  ····················  ····················
E    2766 hCoV-19/India/GJ-GBRC286/2020|EPI_ISL_495024|2020-    ····················  ····················  ····················  ····················  ····················  ····················
E    2767 hCoV-19/India/GJ-GBRC287/2020|EPI_ISL_495025|2020-    ····················  ····················  ····················  ····················  ····················  ····················
E    2768 hCoV-19/India/GJ-GBRC289/2020|EPI_ISL_495026|2020-    ····················  ····················  ····················  ····················  ····················  ····················
E    2769 hCoV-19/India/GJ-GBRC290a/2020|EPI_ISL_495027|2020    ····················  ····················  ····················  ····················  ····················  ···A················
E    2770 hCoV-19/India/GJ-GBRC290b/2020|EPI_ISL_495028|2020    ····················  ····················  ····················  ····················  ····················  ····················
E    2771 hCoV-19/India/GJ-GBRC291/2020|EPI_ISL_495029|2020-    ····················  ····················  ····················  ····················  ····················  ····················
E    2772 hCoV-19/India/GJ-GBRC292a/2020|EPI_ISL_495030|2020    ····················  ····················  ····················  ····················  ····················  ····················
E    2773 hCoV-19/India/GJ-GBRC292b/2020|EPI_ISL_495031|2020    ····················  ····················  ····················  ····················  ····················  ····················
E    2774 hCoV-19/India/GJ-GBRC293a/2020|EPI_ISL_495032|2020    ····················  ····················  ····················  ····················  ····················  ····················
E    2775 hCoV-19/India/GJ-GBRC293b/2020|EPI_ISL_495033|2020    ····················  ····················  ····················  ····················  ····················  ···A················
E    2776 hCoV-19/India/GJ-GBRC294/2020|EPI_ISL_495034|2020-    ····················  ····················  ····················  ····················  ····················  ····················
E    2777 hCoV-19/India/GJ-GBRC295/2020|EPI_ISL_495035|2020-    ····················  ····················  ····················  ····················  ····················  ····················
E    2778 hCoV-19/India/GJ-GBRC296a/2020|EPI_ISL_495036|2020    ····················  ····················  ····················  ····················  ····················  ····················
E    2779 hCoV-19/India/GJ-GBRC296b/2020|EPI_ISL_495037|2020    ····················  ····················  ····················  ····················  ····················  ····················
E    2780 hCoV-19/India/GJ-GBRC297a/2020|EPI_ISL_495038|2020    ····················  ····················  ····················  ····················  ····················  ····················
E    2781 hCoV-19/India/GJ-GBRC297b/2020|EPI_ISL_495039|2020    ····················  ····················  ····················  ····················  ····················  ····················
E    2782 hCoV-19/India/GJ-GBRC298a/2020|EPI_ISL_495040|2020    ····················  ····················  ····················  ····················  ····················  ····················
E    2783 hCoV-19/India/GJ-GBRC298b/2020|EPI_ISL_495041|2020    ····················  ····················  ····················  ····················  ····················  ···A················
E    2784 hCoV-19/India/GJ-GBRC299a/2020|EPI_ISL_495042|2020    ····················  ····················  ····················  ····················  ····················  ····················
E    2785 hCoV-19/India/GJ-GBRC299b/2020|EPI_ISL_495043|2020    ····················  ·A··················  ····················  ····················  ····················  ···A················
E    2786 hCoV-19/India/GJ-GBRC300a/2020|EPI_ISL_495044|2020    ····················  ·A··················  ····················  ····················  ····················  ···A················
E    2787 hCoV-19/India/GJ-GBRC300b/2020|EPI_ISL_495045|2020    ····················  ·A··················  ····················  ····················  ····················  ···A················
E    2788 hCoV-19/India/GJ-GBRC301a/2020|EPI_ISL_495046|2020    ····················  ····················  ····················  ····················  ····················  ····················
E    2789 hCoV-19/India/GJ-GBRC301b/2020|EPI_ISL_495047|2020    ····················  ····················  ····················  ····················  ····················  ····················
E    2790 hCoV-19/India/GJ-GBRC302a/2020|EPI_ISL_495048|2020    ····················  ····················  ····················  ····················  ····················  ···A················
E    2791 hCoV-19/India/GJ-GBRC302b/2020|EPI_ISL_495049|2020    ····················  ·A··················  ····················  ····················  ····················  ···A················
E    2792 hCoV-19/India/GJ-GBRC303a/2020|EPI_ISL_495050|2020    ····················  ·A··················  ····················  ····················  ····················  ···A················
E    2793 hCoV-19/India/GJ-GBRC303b/2020|EPI_ISL_495051|2020    ····················  ····················  ····················  ····················  ····················  ···A················
E    2794 hCoV-19/India/GJ-GBRC304a/2020|EPI_ISL_495052|2020    ····················  ····················  ····················  ····················  ····················  ····················
E    2795 hCoV-19/India/GJ-GBRC304b/2020|EPI_ISL_495053|2020    ····················  ····················  ····················  ····················  ····················  ···A················
E    2796 hCoV-19/India/GJ-GBRC305a/2020|EPI_ISL_495054|2020    ····················  ····················  ····················  ····················  ····················  ····················
E    2797 hCoV-19/India/GJ-GBRC305b/2020|EPI_ISL_495055|2020    ····················  ····················  ····················  ····················  ····················  ···A················
E    2798 hCoV-19/India/GJ-GBRC306a/2020|EPI_ISL_495056|2020    ····················  ·A··················  ····················  ····················  ····················  ···A················
E    2799 hCoV-19/India/GJ-GBRC306b/2020|EPI_ISL_495057|2020    ····················  ·A··················  ····················  ····················  ····················  ···A················
E    2800 hCoV-19/India/GJ-GBRC307a/2020|EPI_ISL_495058|2020    ····················  ····················  ····················  ····················  ····················  ····················
E    2801 hCoV-19/India/GJ-GBRC307b/2020|EPI_ISL_495059|2020    ····················  ····················  ····················  ····················  ····················  ···A················
E    2802 hCoV-19/India/GJ-GBRC308/2020|EPI_ISL_495060|2020-    ····················  ····················  ····················  ····················  ····················  ····················
E    2803 hCoV-19/India/GJ-GBRC309a/2020|EPI_ISL_495061|2020    ····················  ····················  ····················  ····················  ····················  ····················
E    2804 hCoV-19/India/GJ-GBRC309b/2020|EPI_ISL_495062|2020    ····················  ····················  ····················  ····················  ····················  ····················
E    2805 hCoV-19/India/GJ-GBRC310a/2020|EPI_ISL_495063|2020    ····················  ·A··················  ····················  ····················  ····················  ···A················
E    2806 hCoV-19/India/GJ-GBRC310b/2020|EPI_ISL_495064|2020    ····················  ·A··················  ····················  ····················  ····················  ···A················
E    2807 hCoV-19/India/GJ-GBRC311a/2020|EPI_ISL_495065|2020    ····················  ····················  ····················  ····················  ····················  ····················
E    2808 hCoV-19/India/GJ-GBRC311b/2020|EPI_ISL_495066|2020    ····················  ····················  ····················  ····················  ····················  ···A················
```

| | | | | | | | | | |
|---|---|---|---|---|---|---|---|---|---|
| E | 2809 | hCoV-19/India/GJ-GBRC312a/2020\|EPI_ISL_495067\|2020 | ···················· | ···················· | ···················· | ···················· | ···················· | ···················· | |
| E | 2810 | hCoV-19/England/WSFT-25C1950/2020\|EPI_ISL_495090\|2 | ···················· | ·············A······ | ···················· | ···················· | ···················· | ···················· | |
| E | 2811 | hCoV-19/England/WSFT-25C1A8A/2020\|EPI_ISL_495093\|2 | ···················· | ···················· | ···················· | ···················· | ···················· | ···················· | |
| E | 2812 | hCoV-19/England/WSFT-25C1659/2020\|EPI_ISL_495095\|2 | ···················· | ···················· | ···················· | ···················· | ···················· | ···················· | |
| E | 2813 | hCoV-19/England/WSFT-25C16A4/2020\|EPI_ISL_495096\|2 | ···················· | ···················· | ···················· | ···················· | ···················· | ···················· | |
| E | 2814 | hCoV-19/England/BRIS-1854776/2020\|EPI_ISL_495097\|2 | ···················· | ···················· | ···················· | ···················· | ···················· | ···················· | |
| E | 2815 | hCoV-19/England/BRIS-1331E2/2020\|EPI_ISL_495098\|20 | ···················· | ···················· | ···················· | ···················· | ···················· | ···················· | |
| E | 2816 | hCoV-19/England/BRIS-1333BF/2020\|EPI_ISL_495099\|20 | ···················· | ···················· | ···················· | ···················· | ···················· | ···················· | |
| E | 2817 | hCoV-19/Italy/FVG-ICGEB-atr21/2020\|EPI_ISL_479790\| | ···················· | ···················· | ···················· | ···················· | ···················· | ···················· | |
| E | 2818 | hCoV-19/Italy/FVG-ICGEB-S26/2020\|EPI_ISL_479791\|20 | ···················· | ···················· | ···················· | ···················· | ···················· | ···················· | |
| E | 2819 | hCoV-19/Japan/PG-0003/2020\|EPI_ISL_479792\|2020-02 | ···················· | ···················· | ···················· | ···················· | ···················· | ···················· | |
| E | 2820 | hCoV-19/Japan/PG-0004/2020\|EPI_ISL_479793\|2020-02 | ···················· | ···················· | ···················· | ···················· | ···················· | ···················· | |
| E | 2821 | hCoV-19/Japan/PG-0005/2020\|EPI_ISL_479794\|2020-02 | ···················· | ···················· | ···················· | ···················· | ···················· | ···················· | |
| E | 2822 | hCoV-19/Japan/PG-0006/2020\|EPI_ISL_479795\|2020-02 | ···················· | ···················· | ···················· | ···················· | ···················· | ···················· | |
| E | 2823 | hCoV-19/Japan/PG-0009/2020\|EPI_ISL_479796\|2020-02 | ···················· | ···················· | ···················· | ···················· | ···················· | ···················· | |
| E | 2824 | hCoV-19/Japan/PG-0010/2020\|EPI_ISL_479797\|2020-02- | ···················· | ···················· | ···················· | ···················· | ···················· | ···················· | |
| E | 2825 | hCoV-19/Japan/PG-0011/2020\|EPI_ISL_479798\|2020-02- | ···················· | ···················· | ···················· | ···················· | ···················· | ···················· | |
| E | 2826 | hCoV-19/Japan/PG-0015/2020\|EPI_ISL_479799\|2020-01- | ···················· | ···················· | ···················· | ···················· | ···················· | ···················· | |
| E | 2827 | hCoV-19/Japan/PG-0312/2020\|EPI_ISL_479987\|2020-03- | ···················· | ···················· | ···················· | ···················· | ···················· | ···················· | |
| E | 2828 | hCoV-19/Japan/PG-0313/2020\|EPI_ISL_479988\|2020-03- | ···················· | ···················· | ···················· | ···················· | ···················· | ···················· | |
| E | 2829 | hCoV-19/Japan/PG-0314/2020\|EPI_ISL_479989\|2020-03- | ···················· | ···················· | ···················· | ···················· | ···················· | ···················· | |
| E | 2830 | hCoV-19/Japan/PG-0315/2020\|EPI_ISL_479990\|2020-03- | ···················· | ···················· | ···················· | ···················· | ···················· | ···················· | |
| E | 2831 | hCoV-19/Japan/PG-0316/2020\|EPI_ISL_479991\|2020-03- | ···················· | ···················· | ···················· | ···················· | ···················· | ···················· | |
| E | 2832 | hCoV-19/Japan/PG-0317/2020\|EPI_ISL_479992\|2020-03- | ···················· | ···················· | ···················· | ···················· | ···················· | ···················· | |
| E | 2833 | hCoV-19/Japan/PG-0318/2020\|EPI_ISL_479993\|2020-03- | ···················· | ···················· | ···················· | ···················· | ···················· | ···················· | |
| E | 2834 | hCoV-19/Japan/PG-0319/2020\|EPI_ISL_479994\|2020-03- | ···················· | ···················· | ···················· | ···················· | ···················· | ···················· | |
| E | 2835 | hCoV-19/Japan/PG-0320/2020\|EPI_ISL_479995\|2020-03- | ···················· | ···················· | ···················· | ···················· | ···················· | ···················· | |
| E | 2836 | hCoV-19/Japan/PG-0321/2020\|EPI_ISL_479996\|2020-03- | ···················· | ···················· | ···················· | ···················· | ···················· | ···················· | |
| E | 2837 | hCoV-19/Japan/PG-0322/2020\|EPI_ISL_479997\|2020-02- | ···················· | ···················· | ···················· | ···················· | ···················· | ···················· | |
| E | 2838 | hCoV-19/Japan/PG-0323/2020\|EPI_ISL_479998\|2020-02- | ···················· | ···················· | ···················· | ···················· | ···················· | ···················· | |
| E | 2839 | hCoV-19/Japan/PG-0324/2020\|EPI_ISL_479999\|2020-03- | ···················· | ···················· | ···················· | ···················· | ···················· | ···················· | |
| E | 2841 | hCoV-19/Australia/VIC2164/2020\|EPI_ISL_480771\|2020 | ···················· | ···················· | ···················· | ···················· | NNN················· | ···················· | 1 |
| E | 2842 | hCoV-19/Australia/VIC2165/2020\|EPI_ISL_480772\|2020 | ···················· | ···················· | ···················· | ···················· | ···················· | ···················· | |
| E | 2845 | hCoV-19/Australia/VIC2163/2020\|EPI_ISL_480775\|2020 | ···················· | ···················· | ···················· | ···················· | ···················· | ···················· | |
| E | 2848 | hCoV-19/Australia/SA01/2020\|EPI_ISL_480778\|2020-01 | ···················· | ···················· | ···················· | ···················· | ···················· | ···················· | |
| E | 2849 | hCoV-19/Australia/SA02/2020\|EPI_ISL_480779\|2020-01 | ···················· | ···················· | ···················· | ···················· | ···················· | ···················· | |
| E | 2850 | hCoV-19/Australia/SA03/2020\|EPI_ISL_480780\|2020-01 | ···················· | ···················· | ···················· | ···················· | ···················· | ···················· | |
| E | 2851 | hCoV-19/Australia/SA04/2020\|EPI_ISL_480781\|2020-01 | ···················· | ···················· | ···················· | ···················· | ···················· | ···················· | |
| E | 2852 | hCoV-19/Senegal/1683/2020\|EPI_ISL_480783\|2020-03-3 | ···················· | ···················· | ···················· | ···················· | ···················· | ···················· | |
| E | 2853 | hCoV-19/USA/NY-CDC-6683/2020\|EPI_ISL_480784\|2020-0 | ···················· | ···················· | ···················· | ···················· | ···················· | ···················· | |
| E | 2854 | hCoV-19/USA/NY-CDC-6684/2020\|EPI_ISL_480785\|2020-0 | ···················· | ···················· | ···················· | ···················· | ···················· | ···················· | |
| E | 2855 | hCoV-19/Senegal/1749/2020\|EPI_ISL_480787\|2020-03-3 | ···················· | ···················· | ···················· | ···················· | ···················· | ···················· | |
| E | 2856 | hCoV-19/Senegal/1966/2020\|EPI_ISL_480789\|2020-04-0 | ···················· | ···················· | ···················· | ···················· | ···················· | ···················· | |
| E | 2857 | hCoV-19/USA/FL-BPHL-0140/2020\|EPI_ISL_480791\|2020- | ···················· | ···················· | ···················· | ···················· | ···················· | ···················· | |
| E | 2858 | hCoV-19/USA/FL-BPHL-0145/2020\|EPI_ISL_480796\|2020- | ···················· | ···················· | ···················· | ···················· | ···················· | ···················· | |
| E | 2877 | hCoV-19/Japan/TK-YE615667/2020\|EPI_ISL_486900\|2020 | ···················· | ···················· | ···················· | ···················· | ···················· | ···················· | |
| E | 2878 | hCoV-19/Japan/TK-YE614866/2020\|EPI_ISL_486901\|2020 | ···················· | ···················· | ···················· | ···················· | ···················· | ···················· | |
| E | 2879 | hCoV-19/Japan/TK-YE615178/2020\|EPI_ISL_486902\|2020 | ···················· | ···················· | ···················· | ···················· | ···················· | ···················· | |
| E | 2880 | hCoV-19/Japan/TK-YE614327/2020\|EPI_ISL_486903\|2020 | ···················· | ···················· | ···················· | ···················· | ···················· | ···················· | |
| E | 2881 | hCoV-19/Japan/TK-YE614862/2020\|EPI_ISL_486904\|2020 | ···················· | ···················· | ···················· | ···················· | ···················· | ···················· | |
| E | 2882 | hCoV-19/Japan/TK-YE615220/2020\|EPI_ISL_486905\|2020 | ···················· | ···················· | ···················· | ···················· | ···················· | ···················· | |
| E | 2883 | hCoV-19/Japan/TK-YE615221/2020\|EPI_ISL_486906\|2020 | ···················· | ···················· | ···················· | ···················· | ···················· | ···················· | |
| E | 2884 | hCoV-19/Japan/TK-YE614903/2020\|EPI_ISL_486907\|2020 | ···················· | ···················· | ···················· | ···················· | ···················· | ···················· | |
| E | 2885 | hCoV-19/Japan/TK-YE615406/2020\|EPI_ISL_486908\|2020 | ···················· | ···················· | ···················· | ···················· | ···················· | ···················· | |
| E | 2886 | hCoV-19/Japan/TK-Y17873/2020\|EPI_ISL_486909\|2020-0 | ···················· | ···················· | ···················· | ···················· | ···················· | ···················· | |
| E | 2887 | hCoV-19/England/NORT-2921D1/2020\|EPI_ISL_489106\|20 | ···················· | ···················· | ···················· | ···················· | ···················· | ···················· | |
| E | 2888 | hCoV-19/England/NORT-291DFC/2020\|EPI_ISL_489107\|20 | ···················· | ···················· | ···················· | ···················· | ···················· | ···················· | |
| E | 2889 | hCoV-19/England/NORT-29260F/2020\|EPI_ISL_489108\|20 | ···················· | ···················· | ···················· | ···················· | ···················· | ···················· | |
| E | 2890 | hCoV-19/England/NORT-291D56/2020\|EPI_ISL_489109\|20 | ···················· | ···················· | ···················· | ···················· | ···················· | ···················· | |
| E | 2891 | hCoV-19/England/NORT-292757/2020\|EPI_ISL_489110\|20 | ···················· | ···················· | ···················· | ···················· | ···················· | ···················· | |
| E | 2892 | hCoV-19/England/NORT-292E07/2020\|EPI_ISL_489113\|20 | ···················· | ···················· | ···················· | ···················· | ···················· | ···················· | |
| E | 2893 | hCoV-19/England/NORT-291E17/2020\|EPI_ISL_489114\|20 | ···················· | ···················· | ···················· | ···················· | ···················· | ···················· | |
| E | 2894 | hCoV-19/England/NORT-29271B/2020\|EPI_ISL_489117\|20 | ···················· | ···················· | ···················· | ···················· | ···················· | ···················· | |
| E | 2895 | hCoV-19/England/NORT-291C95/2020\|EPI_ISL_489119\|20 | ···················· | ···················· | ···················· | ···················· | ···················· | ···················· | |
| E | 2896 | hCoV-19/England/BRIS-131937/2020\|EPI_ISL_489120\|20 | ···················· | ···················· | ···················· | ···················· | ···················· | ···················· | |
| E | 2897 | hCoV-19/England/NORT-292854/2020\|EPI_ISL_489121\|20 | ···················· | ···················· | ···················· | ···················· | ···················· | ···················· | |
| E | 2898 | hCoV-19/England/NORT-292186/2020\|EPI_ISL_489122\|20 | ···················· | ···················· | ···················· | ···················· | ···················· | ···················· | |
| E | 2899 | hCoV-19/England/NORT-291DA1/2020\|EPI_ISL_489123\|20 | ···················· | ···················· | ···················· | ···················· | ···················· | ···················· | |
| E | 2900 | hCoV-19/England/NORT-29262D/2020\|EPI_ISL_489124\|20 | ···················· | ···················· | ···················· | ···················· | ···················· | ···················· | |
| E | 2901 | hCoV-19/England/NORT-2922ED/2020\|EPI_ISL_489125\|20 | ···················· | ···················· | ···················· | ···················· | ···················· | ···················· | |
| E | 2902 | hCoV-19/England/NORT-2923AE/2020\|EPI_ISL_489126\|20 | ···················· | ···················· | ···················· | ···················· | ···················· | ···················· | |
| E | 2903 | hCoV-19/England/NORT-2925E4/2020\|EPI_ISL_489127\|20 | ···················· | ···················· | ···················· | ···················· | ···················· | ···················· | |
| E | 2904 | hCoV-19/England/NORT-291CD1/2020\|EPI_ISL_489128\|20 | ···················· | ···················· | ···················· | ···················· | ···················· | ···················· | |
| E | 2905 | hCoV-19/England/NORT-292BF1/2020\|EPI_ISL_489129\|20 | ···················· | ···················· | ···················· | ···················· | ···················· | ···················· | |
| E | 2906 | hCoV-19/England/NORT-291D92/2020\|EPI_ISL_489131\|20 | ···················· | ···················· | ···················· | ···················· | ···················· | ···················· | |
| E | 2907 | hCoV-19/England/NORT-291DED/2020\|EPI_ISL_489132\|20 | ···················· | ···················· | ···················· | ···················· | ···················· | ···················· | |
| E | 2908 | hCoV-19/England/NORT-292353/2020\|EPI_ISL_489133\|20 | ···················· | ···················· | ···················· | ···················· | ···················· | ···················· | |
| E | 2909 | hCoV-19/England/NORT-29203E/2020\|EPI_ISL_489135\|20 | ···················· | ···················· | ···················· | ···················· | ···················· | ···················· | |
| E | 2910 | hCoV-19/England/NORT-292D46/2020\|EPI_ISL_489137\|20 | ···················· | ···················· | ···················· | ···················· | ···················· | ···················· | |
| E | 2911 | hCoV-19/England/NORT-291D47/2020\|EPI_ISL_489138\|20 | ···················· | ···················· | ···················· | ···················· | ···················· | ···················· | |

E 2912 hCoV-19/England/NORT-2921C2/2020|EPI_ISL_489139|20 .................... .................... .................... .................... .................... ....................
E 2913 hCoV-19/England/NORT-292E16/2020|EPI_ISL_489140|20 .................... .................... .................... .................... .................... ....................
E 2914 hCoV-19/England/NORT-292DFB/2020|EPI_ISL_489141|20 .................... .................... .................... .................... .................... ....................
E 2915 hCoV-19/England/NORT-29298E/2020|EPI_ISL_489142|20 .................... .................... .................... .................... .................... ....................
E 2916 hCoV-19/England/NORT-292441/2020|EPI_ISL_489143|20 .................... .................... .................... .................... .................... ....................
E 2917 hCoV-19/England/NORT-292414/2020|EPI_ISL_489144|20 .................... .................... .................... .................... .................... ....................
E 2918 hCoV-19/England/NORT-291D83/2020|EPI_ISL_489146|20 .................... .................... .................... .................... .................... ....................
E 2919 hCoV-19/England/NORT-292177/2020|EPI_ISL_489149|20 .................... .................... .................... .................... .................... ....................
E 2920 hCoV-19/Northern Ireland/NIRE-FED95/2020|EPI_ISL_4 .................... .................... .................... .................... .................... ....................
E 2921 hCoV-19/Northern Ireland/NIRE-10328E/2020|EPI_ISL_ .................... .................... .................... .................... .................... ....................
E 2922 hCoV-19/Northern Ireland/NIRE-FD0CA/2020|EPI_ISL_4 .................... .................... .................... .................... .................... ....................
E 2923 hCoV-19/Northern Ireland/NIRE-FCD2E/2020|EPI_ISL_4 .................... .................... .................... .................... .................... ....................
E 2924 hCoV-19/Northern Ireland/NIRE-1036BF/2020|EPI_ISL_ .................... .................... .................... .................... .................... ....................
E 2925 hCoV-19/Northern Ireland/NIRE-100139/2020|EPI_ISL_ .................... .................... .................... .................... .................... ....................
E 2926 hCoV-19/Northern Ireland/NIRE-FD288/2020|EPI_ISL_4 .................... .................... .................... .................... .................... ....................
E 2927 hCoV-19/Northern Ireland/NIRE-102A5A/2020|EPI_ISL_ .................... .................... .................... .................... .................... ....................
E 2928 hCoV-19/Northern Ireland/NIRE-103AA4/2020|EPI_ISL_ .................... .................... .................... .................... .................... ....................
E 2929 hCoV-19/Northern Ireland/NIRE-FCDF1/2020|EPI_ISL_4 .................... .................... .................... .................... .................... ....................
E 2930 hCoV-19/Northern Ireland/NIRE-10385F/2020|EPI_ISL_ .................... .................... .................... .................... .................... ....................
E 2931 hCoV-19/Northern Ireland/NIRE-FD886/2020|EPI_ISL_4 .................... .................... .................... .................... .................... ....................
E 2932 hCoV-19/Northern Ireland/NIRE-FE7F1/2020|EPI_ISL_4 .................... .................... .................... .................... .................... ....................
E 2933 hCoV-19/Northern Ireland/NIRE-102AB4/2020|EPI_ISL_ .................... .................... .................... .................... .................... ....................
E 2934 hCoV-19/Northern Ireland/NIRE-10195E/2020|EPI_ISL_ .................... .................... .................... .................... .................... ....................
E 2935 hCoV-19/Northern Ireland/NIRE-10329D/2020|EPI_ISL_ .................... .................... .................... .................... .................... ....................
E 2936 hCoV-19/Northern Ireland/NIRE-10345B/2020|EPI_ISL_ .................... .................... .................... .................... .................... ....................
E 2937 hCoV-19/Northern Ireland/NIRE-103549/2020|EPI_ISL_ .................... .................... .................... .................... .................... ....................
E 2938 hCoV-19/Northern Ireland/NIRE-103118/2020|EPI_ISL_ .................... .................... .................... .................... .................... ....................
E 2939 hCoV-19/Northern Ireland/NIRE-103A0E/2020|EPI_ISL_ .................... .................... .................... .................... .................... ....................
E 2940 hCoV-19/Northern Ireland/NIRE-1034C4/2020|EPI_ISL_ .................... .................... .................... .................... .................... ....................
E 2941 hCoV-19/Northern Ireland/NIRE-FCED0/2020|EPI_ISL_4 .................... .................... .................... .................... .................... ....................
E 2942 hCoV-19/Northern Ireland/NIRE-FD0F7/2020|EPI_ISL_4 .................... .................... .................... .................... .................... ....................
E 2943 hCoV-19/Northern Ireland/NIRE-FD200/2020|EPI_ISL_4 .................... .................... .................... .................... .................... ....................
E 2944 hCoV-19/Northern Ireland/NIRE-1001EE/2020|EPI_ISL_ .................... .................... .................... .................... .................... ....................
E 2945 hCoV-19/Northern Ireland/NIRE-1003F7/2020|EPI_ISL_ .................... .................... .................... .................... .................... ....................
E 2946 hCoV-19/Northern Ireland/NIRE-103136/2020|EPI_ISL_ .................... .................... .................... .................... .................... ....................
E 2947 hCoV-19/Northern Ireland/NIRE-FFFE9/2020|EPI_ISL_4 .................... .................... .................... .................... .................... ....................
E 2948 hCoV-19/Malaysia/IMR_WC99045/2020|EPI_ISL_490100|2 .................... .................... .................... .................... .................... ....................
E 2949 hCoV-19/Malaysia/IMR_WC90685/2020|EPI_ISL_490101|2 .................... .................... .................... .................... .................... ....................
E 2950 hCoV-19/Malaysia/IMR_WC94764/2020|EPI_ISL_490103|2 .................... .................... .................... .................... .................... ....................
E 2951 hCoV-19/India/UN-7044/2020|EPI_ISL_490106|2020 .................... .................... .................... .................... .................... ....................
E 2952 hCoV-19/Bangladesh/BCSIR-NILMRC-246/2020|EPI_ISL_4 .................... .................... .................... .................... .................... ....................
E 2953 hCoV-19/Bangladesh/BCSIR-NILMRC-247/2020|EPI_ISL_4 .................... .................... .................... .................... .................... ....................
E 2954 hCoV-19/Bangladesh/BCSIR-NILMRC-248/2020|EPI_ISL_4 .................... .................... .................... .................... .................... ....................
E 2955 hCoV-19/Czech Republic/NRL_3969/2020|EPI_ISL_49011 .................... .................... .................... .................... .................... ....................
E 2956 hCoV-19/Bangladesh/BCSIR-NILMRC-249/2020|EPI_ISL_4 .................... .................... .................... .................... .................... ....................
E 2957 hCoV-19/Bangladesh/BCSIR-NILMRC-250/2020|EPI_ISL_4 .................... .................... .................... .................... .................... ....................
E 2958 hCoV-19/Bangladesh/BCSIR-NILMRC-253/2020|EPI_ISL_4 .................... .................... .................... .................... .................... ....................
E 2959 hCoV-19/Bangladesh/BCSIR-NILMRC-256/2020|EPI_ISL_4 .................... .................... .................... .................... .................... ....................
E 2960 hCoV-19/Bangladesh/BCSIR-NILMRC-255/2020|EPI_ISL_4 .................... .................... .................... .................... .................... ....................
E 2961 hCoV-19/Bangladesh/BCSIR-NILMRC-257/2020|EPI_ISL_4 .................... .A.................. .................... .................... .................... ..A.................
E 2962 hCoV-19/USA/WI-UW-747/2020|EPI_ISL_491420|2020-07- .................... .................... .................... .................... .................... ....................
E 2963 hCoV-19/USA/WI-UW-748/2020|EPI_ISL_491421|2020-06- .................... .................... .................... .................... .................... ....................
E 2964 hCoV-19/Peru/LIM-INS-021/2020|EPI_ISL_491427|2020- .................... .................... .................... .................... .................... ....................
E 2965 hCoV-19/Peru/LIM-INS-022/2020|EPI_ISL_491428|2020- .................... .................... .................... .................... .................... ....................
E 2966 hCoV-19/Peru/LIM-INS-024/2020|EPI_ISL_491429|2020- .................... .................... .................... .................... .................... ....................
E 2967 hCoV-19/Peru/LIM-INS-025/2020|EPI_ISL_491430|2020- .................... .................... .................... .................... .................... ....................
E 2968 hCoV-19/Peru/ARE-INS-026/2020|EPI_ISL_491431|2020- .................... .................... .................... .................... .................... ....................
E 2969 hCoV-19/Peru/LIM-INS-028/2020|EPI_ISL_491432|2020- .................... .................... .................... .................... .................... ....................
E 2970 hCoV-19/Peru/LIM-INS-029/2020|EPI_ISL_491433|2020- .................... .................... .................... .................... .................... ....................
E 2971 hCoV-19/Peru/LIM-INS-030/2020|EPI_ISL_491434|2020- .................... .................... .................... .................... .................... ....................
E 2972 hCoV-19/Peru/LIM-INS-031/2020|EPI_ISL_491435|2020- .................... .................... .................... .................... .................... ....................
E 2973 hCoV-19/Peru/LIM-INS-032/2020|EPI_ISL_491436|2020- .................... .................... .................... .................... .................... ....................
E 2974 hCoV-19/Costa Rica/INC-0025/2020|EPI_ISL_491437|20 .................... .................... .................... .................... .................... ....................
E 2975 hCoV-19/Costa Rica/INC-0026/2020|EPI_ISL_491438|20 .................... .................... .................... .................... .................... ....................
E 2976 hCoV-19/Costa Rica/INC-0027/2020|EPI_ISL_491439|20 .................... .................... .................... .................... .................... ....................
E 2977 hCoV-19/Costa Rica/INC-0028/2020|EPI_ISL_491440|20 .................... .................... .................... .................... .................... ....................
E 2978 hCoV-19/Costa Rica/INC-0029/2020|EPI_ISL_491441|20 .................... .................... .................... .................... .................... ....................
E 2979 hCoV-19/Costa Rica/INC-0030/2020|EPI_ISL_491442|20 .................... .................... .................... .................... .................... ....................
E 2980 hCoV-19/Costa Rica/INC-0031/2020|EPI_ISL_491443|20 .................... .................... .................... .................... .................... ....................
E 2981 hCoV-19/Costa Rica/INC-0032/2020|EPI_ISL_491444|20 .................... .................... .................... .................... .................... ....................
E 2982 hCoV-19/Costa Rica/INC-0033/2020|EPI_ISL_491445|20 .................... .................... .................... .................... .................... ....................
E 2983 hCoV-19/Costa Rica/INC-0034/2020|EPI_ISL_491446|20 .................... .................... .................... .................... .................... ....................
E 2984 hCoV-19/Costa Rica/INC-0035/2020|EPI_ISL_491447|20 .................... .................... .................... .................... .................... ....................
E 2985 hCoV-19/Costa Rica/INC-0037/2020|EPI_ISL_491448|20 .................... .................... .................... .................... .................... ....................
E 2986 hCoV-19/Costa Rica/INC-0038/2020|EPI_ISL_491449|20 .................... .................... .................... .................... .................... ....................
E 2987 hCoV-19/Costa Rica/INC-0039/2020|EPI_ISL_491450|20 .................... .................... .................... .................... .................... ....................
E 2988 hCoV-19/Costa Rica/INC-0040/2020|EPI_ISL_491451|20 .................... .................... .................... .................... .................... ....................
E 2989 hCoV-19/Costa Rica/INC-0041/2020|EPI_ISL_491452|20 .................... .................... .................... .................... .................... ....................
E 2990 hCoV-19/Costa Rica/INC-0042/2020|EPI_ISL_491453|20 .................... .................... .................... .................... .................... ....................
E 2991 hCoV-19/Costa Rica/INC-0043/2020|EPI_ISL_491454|20 .................... .................... .................... .................... .................... ....................

| | | | | | | | | |
|---|---|---|---|---|---|---|---|---|
| E | 2992 hCoV-19/Costa Rica/INC-0044/2020\|EPI_ISL_491455\|20 | .................... | .................... | ..................... | .................... | .................... | .................... | |
| E | 2993 hCoV-19/Costa Rica/INC-0045/2020\|EPI_ISL_491456\|20 | .................... | .................... | ..................... | .................... | .................... | .................... | |
| E | 2994 hCoV-19/Costa Rica/INC-0046/2020\|EPI_ISL_491457\|20 | .................... | .................... | ..................... | .................... | .................... | .................... | |
| E | 2995 hCoV-19/Peru/LIM-INS-033/2020\|EPI_ISL_491458\|2020- | .................... | .................... | ..................... | .................... | .................... | .................... | |
| E | 2996 hCoV-19/Peru/LAM-INS-034/2020\|EPI_ISL_491459\|2020- | .................... | .................... | ..................... | .................... | .................... | .................... | |
| E | 2997 hCoV-19/Peru/LIM-INS-013/2020\|EPI_ISL_491463\|2020- | .................... | .................... | ..................... | .................... | .................... | .................... | |
| E | 2998 hCoV-19/Peru/ICA-INS-039/2020\|EPI_ISL_491464\|2020- | .................... | .................... | ..................... | .................... | .................... | .................... | |
| E | 2999 hCoV-19/Scotland/EDB3913/2020\|EPI_ISL_491494\|2020- | .................... | .................... | ..................... | .................... | .................... | .................... | |
| E | 3000 hCoV-19/Scotland/EDB1400/2020\|EPI_ISL_491495\|2020- | .................... | .................... | ..................... | .................... | .................... | .................... | |
| E | 3001 hCoV-19/Scotland/EDB2167/2020\|EPI_ISL_491496\|2020- | .................... | .................... | ..................... | .................... | .................... | .................... | |
| E | 3002 hCoV-19/Scotland/EDB2571/2020\|EPI_ISL_491497\|2020- | .................... | .A.................. | ..................... | .................... | .................... | .................... | |
| E | 3003 hCoV-19/Scotland/EDB2250/2020\|EPI_ISL_491498\|2020- | .................... | .................... | ..................... | .................... | .................... | .................... | |
| E | 3004 hCoV-19/Scotland/EDB1974/2020\|EPI_ISL_491620\|2020- | .................... | ...................A | ..................... | .................... | .................... | .................... | 1 |
| E | 3005 hCoV-19/Scotland/EDB2370/2020\|EPI_ISL_491621\|2020- | .................... | .................... | ..................... | .................... | .................... | .................... | |
| E | 3006 hCoV-19/Scotland/EDB2485/2020\|EPI_ISL_491622\|2020- | .................... | .................... | ..................... | .................... | .................... | .................... | |
| E | 3007 hCoV-19/Scotland/EDB1849/2020\|EPI_ISL_491623\|2020- | .................... | .................... | ..................... | .................... | .................... | .................... | |
| E | 3008 hCoV-19/Scotland/EDB1484/2020\|EPI_ISL_491624\|2020- | .................... | .................... | ..................... | .................... | .................... | .................... | |
| E | 3009 hCoV-19/Scotland/EDB1899/2020\|EPI_ISL_491625\|2020- | .................... | .................... | ..................... | .................... | .................... | .................... | |
| E | 3010 hCoV-19/Scotland/EDB2473/2020\|EPI_ISL_491626\|2020- | .................... | .................... | ..................... | .................... | .................... | .................... | |
| E | 3011 hCoV-19/Scotland/EDB2470/2020\|EPI_ISL_491627\|2020- | .................... | .................... | ..................... | .................... | .................... | .................... | |
| E | 3012 hCoV-19/Scotland/EDB1835/2020\|EPI_ISL_491628\|2020- | .................... | .................... | ..................... | .................... | .................... | .................... | |
| E | 3013 hCoV-19/Scotland/EDB6735/2020\|EPI_ISL_491630\|2020- | .................... | .................... | ..................... | .................... | .................... | .................... | |
| E | 3014 hCoV-19/Scotland/EDB2520/2020\|EPI_ISL_491631\|2020- | .................... | ...................A | ..................... | .................... | .................... | .................... | 1 |
| E | 3015 hCoV-19/Scotland/EDB1995/2020\|EPI_ISL_491635\|2020- | .................... | .A.................. | ..................... | .................... | .................... | .................... | |
| E | 3016 hCoV-19/Scotland/EDB1394/2020\|EPI_ISL_491636\|2020- | .................... | .A.................. | ..................... | .................... | .................... | .................... | |
| E | 3017 hCoV-19/Scotland/EDB872/2020\|EPI_ISL_491637\|2020-0 | .................... | .A.................. | ..................... | .................... | .................... | .................... | |
| E | 3018 hCoV-19/Scotland/EDB6728/2020\|EPI_ISL_491638\|2020- | .................... | .................... | ..................... | .................... | .................... | .................... | |
| E | 3019 hCoV-19/Scotland/EDB6745/2020\|EPI_ISL_491639\|2020- | .................... | .................... | ..................... | .................... | .................... | .................... | |
| E | 3020 hCoV-19/Scotland/EDB1233/2020\|EPI_ISL_491650\|2020- | .................... | .................... | ..................... | .................... | .................... | .................... | |
| E | 3021 hCoV-19/England/LOND-D899D/2020\|EPI_ISL_492751\|202 | .................... | .................... | ..................... | .................... | .................... | .................... | |
| E | 3022 hCoV-19/England/LOND-D8450/2020\|EPI_ISL_492753\|202 | .................... | .................... | ..................... | .................... | .................... | .................... | |
| E | 3023 hCoV-19/England/LOND-D89AC/2020\|EPI_ISL_492754\|202 | .................... | .................... | ..................... | .................... | .................... | .................... | |
| E | 3024 hCoV-19/England/LOND-D861E/2020\|EPI_ISL_492755\|202 | .................... | .................... | ..................... | .................... | .................... | .................... | |
| E | 3025 hCoV-19/England/LOND-D88BE/2020\|EPI_ISL_492756\|202 | .................... | .................... | ..................... | .................... | .................... | .................... | |
| E | 3026 hCoV-19/England/LOND-D8AE5/2020\|EPI_ISL_492760\|202 | .................... | .................... | ..................... | .................... | .................... | .................... | |
| E | 3027 hCoV-19/England/BRIS-123D0F/2020\|EPI_ISL_492764\|20 | .................... | .................... | ..................... | .................... | .................... | .................... | |
| E | 3028 hCoV-19/England/BRIS-124612/2020\|EPI_ISL_492765\|20 | .................... | .................... | ..................... | .................... | .................... | .................... | |
| E | 3029 hCoV-19/England/BRIS-124050/2020\|EPI_ISL_492766\|20 | .................... | .................... | ..................... | .................... | .................... | .................... | |
| E | 3030 hCoV-19/England/BRIS-123FAF/2020\|EPI_ISL_492767\|20 | .................... | .................... | ..................... | .................... | .................... | .................... | |
| E | 3031 hCoV-19/England/BRIS-123CE4/2020\|EPI_ISL_492768\|20 | .................... | .................... | ..................... | .................... | .................... | .................... | |
| E | 3032 hCoV-19/England/BRIS-12413F/2020\|EPI_ISL_492769\|20 | .................... | .................... | ..................... | .................... | .................... | .................... | |
| E | 3033 hCoV-19/England/BRIS-123D78/2020\|EPI_ISL_492770\|20 | .................... | .................... | ..................... | .................... | .................... | .................... | |
| E | 3034 hCoV-19/England/BRIS-123FCD/2020\|EPI_ISL_492771\|20 | .................... | .................... | ..................... | .................... | .................... | .................... | |
| E | 3035 hCoV-19/England/BRIS-124023/2020\|EPI_ISL_492772\|20 | .................... | .................... | ..................... | .................... | .................... | .................... | |
| E | 3036 hCoV-19/England/BRIS-124463/2020\|EPI_ISL_492774\|20 | .................... | .................... | ..................... | .................... | .................... | .................... | |
| E | 3037 hCoV-19/England/BRIS-1240BA/2020\|EPI_ISL_492775\|20 | .................... | .................... | ..................... | .................... | .................... | .................... | |
| E | 3038 hCoV-19/England/BRIS-1240F6/2020\|EPI_ISL_492776\|20 | .................... | .................... | ..................... | .................... | .................... | .................... | |
| E | 3039 hCoV-19/England/BRIS-123D2D/2020\|EPI_ISL_492777\|20 | .................... | .................... | ..................... | .................... | .................... | .................... | |
| E | 3040 hCoV-19/England/BRIS-124454/2020\|EPI_ISL_492778\|20 | .................... | .................... | ..................... | .................... | .................... | .................... | |
| E | 3041 hCoV-19/England/BRIS-12415D/2020\|EPI_ISL_492780\|20 | .................... | .................... | ..................... | .................... | .................... | .................... | |
| E | 3042 hCoV-19/England/BRIS-123F36/2020\|EPI_ISL_492782\|20 | .................... | .............A...... | ..................... | .................... | .................... | .................... | |
| E | 3043 hCoV-19/England/BRIS-1240C9/2020\|EPI_ISL_492783\|20 | .................... | .................... | ..................... | .................... | .................... | .................... | |
| E | 3044 hCoV-19/England/BRIS-123DA5/2020\|EPI_ISL_492784\|20 | .................... | .................... | ..................... | .................... | .................... | .................... | |
| E | 3045 hCoV-19/England/BRIS-12418A/2020\|EPI_ISL_492785\|20 | .................... | .................... | ..................... | .................... | .................... | .................... | |
| E | 3046 hCoV-19/England/BRIS-1246A9/2020\|EPI_ISL_492787\|20 | .................... | .................... | ..................... | .................... | .................... | .................... | |
| E | 3047 hCoV-19/England/BRIS-123EDF/2020\|EPI_ISL_492788\|20 | .................... | .................... | ..................... | .................... | .................... | .................... | |
| E | 3048 hCoV-19/England/BRIS-124120/2020\|EPI_ISL_492789\|20 | .................... | .................... | ..................... | .................... | .................... | .................... | |
| E | 3049 hCoV-19/England/BRIS-12406F/2020\|EPI_ISL_492790\|20 | .................... | .................... | ..................... | .................... | .................... | .................... | |
| E | 3050 hCoV-19/England/BRIS-12421E/2020\|EPI_ISL_492794\|20 | .................... | .................... | ..................... | .................... | .................... | .................... | |
| E | 3051 hCoV-19/England/BRIS-124032/2020\|EPI_ISL_492795\|20 | .................... | .................... | ..................... | .................... | .................... | .................... | |
| E | 3052 hCoV-19/England/BRIS-123F09/2020\|EPI_ISL_492796\|20 | .................... | .................... | ..................... | .................... | .................... | .................... | |
| E | 3053 hCoV-19/England/BRIS-124041/2020\|EPI_ISL_492797\|20 | .................... | .................... | ..................... | .................... | .................... | .................... | |
| E | 3054 hCoV-19/England/BRIS-124560/2020\|EPI_ISL_492798\|20 | .................... | .................... | ..................... | .................... | .................... | .................... | |
| E | 3055 hCoV-19/England/BRIS-123EFD/2020\|EPI_ISL_492799\|20 | .................... | .................... | ..................... | .................... | .................... | .................... | |
| E | 3056 hCoV-19/England/NORT-28C565/2020\|EPI_ISL_492951\|20 | .................... | .................... | ..................... | .................... | .................... | .................... | |
| E | 3057 hCoV-19/England/NORT-28C240/2020\|EPI_ISL_492952\|20 | .................... | .................... | ..................... | .................... | .................... | .................... | |
| E | 3058 hCoV-19/England/NORT-28B19F/2020\|EPI_ISL_492953\|20 | .................... | .................... | ..................... | .................... | .................... | .................... | |
| E | 3059 hCoV-19/England/NORT-28C459/2020\|EPI_ISL_492954\|20 | .................... | .................... | ..................... | .................... | .................... | .................... | |
| E | 3060 hCoV-19/England/NORT-28AE3F/2020\|EPI_ISL_492955\|20 | .................... | .................... | ..................... | .................... | .................... | .................... | |
| E | 3061 hCoV-19/England/NORT-28AEC6/2020\|EPI_ISL_492956\|20 | .................... | .................... | ..................... | .................... | .................... | .................... | |
| E | 3062 hCoV-19/England/NORT-28C310/2020\|EPI_ISL_492958\|20 | .................... | .................... | ..................... | .................... | .................... | .................... | |
| E | 3063 hCoV-19/England/NORT-28ADE7/2020\|EPI_ISL_492959\|20 | .................... | .................... | ..................... | .................... | .................... | .................... | |
| E | 3064 hCoV-19/England/NORT-28AD14/2020\|EPI_ISL_492962\|20 | .................... | .................... | ..................... | .................... | .................... | .................... | |
| E | 3065 hCoV-19/England/NORT-28ADC9/2020\|EPI_ISL_492963\|20 | .................... | .................... | ..................... | .................... | .................... | .................... | |
| E | 3066 hCoV-19/England/NORT-28B0A1/2020\|EPI_ISL_492964\|20 | .................... | .................... | ..................... | .................... | .................... | .................... | |
| E | 3067 hCoV-19/England/NORT-28AD23/2020\|EPI_ISL_492965\|20 | .................... | .................... | ..................... | .................... | .................... | .................... | |
| E | 3068 hCoV-19/England/NORT-28AD50/2020\|EPI_ISL_492966\|20 | .................... | .................... | ..................... | .................... | .................... | .................... | |
| E | 3069 hCoV-19/England/NORT-28ADF6/2020\|EPI_ISL_492968\|20 | .................... | .................... | ..................... | .................... | .................... | .................... | |
| E | 3070 hCoV-19/England/NORT-28AF2D/2020\|EPI_ISL_492969\|20 | .................... | .................... | ..................... | .................... | .................... | .................... | |
| E | 3071 hCoV-19/Italy/VEN-UniVR-1/2020\|EPI_ISL_492980\|2020 | .................... | .................... | ..................... | .................... | .................... | .................... | |

| | | | | | | | | |
|---|---|---|---|---|---|---|---|---|
| E | 3072 | hCoV-19/Italy/VEN-UniVR-2/2020\|EPI_ISL_492981\|2020 | .................... | .................... | .................... | .................... | .................... | .................... |
| E | 3073 | hCoV-19/Japan/PG-0195/2020\|EPI_ISL_479900\|2020-03- | .................... | .................... | .................... | .................... | .................... | .................... |
| E | 3074 | hCoV-19/Japan/PG-0196/2020\|EPI_ISL_479901\|2020-03- | .................... | .................... | .................... | .................... | .................... | .................... |
| E | 3075 | hCoV-19/Japan/PG-0197/2020\|EPI_ISL_479902\|2020-03- | .................... | .................... | .................... | .................... | .................... | .................... |
| E | 3076 | hCoV-19/Japan/PG-0200/2020\|EPI_ISL_479903\|2020-03- | .................... | .................... | .................... | .................... | .................... | .................... |
| E | 3077 | hCoV-19/Japan/PG-0201/2020\|EPI_ISL_479904\|2020-03- | .................... | .................... | .................... | .................... | .................... | .................... |
| E | 3078 | hCoV-19/Japan/PG-0204/2020\|EPI_ISL_479905\|2020-03- | .................... | .................... | .................... | .................... | .................... | .................... |
| E | 3079 | hCoV-19/Japan/PG-0205/2020\|EPI_ISL_479906\|2020-03- | .................... | .................... | .................... | .................... | .................... | .................... |
| E | 3080 | hCoV-19/Japan/PG-0207/2020\|EPI_ISL_479907\|2020-03- | .................... | .................... | .................... | .................... | .................... | .................... |
| E | 3081 | hCoV-19/Japan/PG-0208/2020\|EPI_ISL_479908\|2020-03- | .................... | .................... | .................... | .................... | .................... | .................... |
| E | 3082 | hCoV-19/Japan/PG-0209/2020\|EPI_ISL_479909\|2020-03- | .................... | .................... | .................... | .................... | .................... | .................... |
| E | 3083 | hCoV-19/Japan/PG-0210/2020\|EPI_ISL_479910\|2020-03- | .................... | .................... | .................... | .................... | .................... | .................... |
| E | 3084 | hCoV-19/Japan/PG-0211/2020\|EPI_ISL_479911\|2020-03- | .................... | .................... | .................... | .................... | .................... | .................... |
| E | 3085 | hCoV-19/Japan/PG-0212/2020\|EPI_ISL_479912\|2020-03- | .................... | .................... | .................... | .................... | .................... | .................... |
| E | 3086 | hCoV-19/Japan/PG-0213/2020\|EPI_ISL_479913\|2020-02- | .................... | .................... | .................... | .................... | .................... | .................... |
| E | 3087 | hCoV-19/Japan/PG-0214/2020\|EPI_ISL_479914\|2020-03- | .................... | .................... | .................... | .................... | .................... | .................... |
| E | 3088 | hCoV-19/Japan/PG-0215/2020\|EPI_ISL_479915\|2020-03- | .................... | .................... | .................... | .................... | .................... | .................... |
| E | 3089 | hCoV-19/Japan/PG-0216/2020\|EPI_ISL_479916\|2020-03- | .................... | .................... | .................... | .................... | .................... | .................... |
| E | 3090 | hCoV-19/Japan/PG-0217/2020\|EPI_ISL_479917\|2020-03- | .................... | .................... | .................... | .................... | .................... | .................... |
| E | 3091 | hCoV-19/Japan/PG-0218/2020\|EPI_ISL_479918\|2020-03- | .................... | .................... | .................... | .................... | .................... | .................... |
| E | 3092 | hCoV-19/Japan/PG-0219/2020\|EPI_ISL_479919\|2020-03- | .................... | .................... | .................... | .................... | .................... | .................... |
| E | 3093 | hCoV-19/Japan/PG-0221/2020\|EPI_ISL_479920\|2020-03- | .................... | .................... | .................... | .................... | .................... | .................... |
| E | 3094 | hCoV-19/Japan/PG-0259/2020\|EPI_ISL_479943\|2020-03- | .................... | .................... | .................... | .................... | .................... | .................... |
| E | 3095 | hCoV-19/Japan/PG-0260/2020\|EPI_ISL_479944\|2020-03 | .................... | .................... | .................... | .................... | .................... | .................... |
| E | 3096 | hCoV-19/Japan/PG-0261/2020\|EPI_ISL_479945\|2020-03 | .................... | .................... | .................... | .................... | .................... | .................... |
| E | 3097 | hCoV-19/Japan/PG-0262/2020\|EPI_ISL_479946\|2020-03 | .................... | .................... | .................... | .................... | .................... | .................... |
| E | 3098 | hCoV-19/Japan/PG-0263/2020\|EPI_ISL_479947\|2020-03 | .................... | .................... | .................... | .................... | .................... | .................... |
| E | 3099 | hCoV-19/Japan/PG-0264/2020\|EPI_ISL_479948\|2020-03 | .................... | .................... | .................... | .................... | .................... | .................... |
| E | 3100 | hCoV-19/Japan/PG-0265/2020\|EPI_ISL_479949\|2020-03 | .................... | .................... | .................... | .................... | .................... | .................... |
| E | 3101 | hCoV-19/Japan/PG-0267/2020\|EPI_ISL_479950\|2020-03 | .................... | .................... | .................... | .................... | .................... | .................... |
| E | 3102 | hCoV-19/Japan/PG-0268/2020\|EPI_ISL_479951\|2020-03 | .................... | .................... | .................... | .................... | .................... | .................... |
| E | 3103 | hCoV-19/Japan/PG-0269/2020\|EPI_ISL_479952\|2020-03 | .................... | .................... | .................... | ..........C......... | .................... | .................... |
| E | 3104 | hCoV-19/Japan/PG-0271/2020\|EPI_ISL_479953\|2020-03 | .................... | .................... | .................... | .................... | .................... | .................... |
| E | 3105 | hCoV-19/Japan/PG-0272/2020\|EPI_ISL_479954\|2020-03 | .................... | .................... | .................... | .................... | .................... | .................... |
| E | 3106 | hCoV-19/Japan/PG-0274/2020\|EPI_ISL_479955\|2020-03 | .................... | .................... | .................... | .................... | .................... | .................... |
| E | 3107 | hCoV-19/Japan/PG-0275/2020\|EPI_ISL_479956\|2020-03 | .................... | .................... | .................... | .................... | .................... | .................... |
| E | 3108 | hCoV-19/Japan/PG-0276/2020\|EPI_ISL_479957\|2020-03 | .................... | .................... | .................... | .................... | .................... | .................... |
| E | 3109 | hCoV-19/Japan/PG-0277/2020\|EPI_ISL_479958\|2020-03 | .................... | .................... | .................... | .................... | .................... | .................... |
| E | 3110 | hCoV-19/Japan/PG-0279/2020\|EPI_ISL_479959\|2020-03- | .................... | .................... | .................... | .................... | .................... | .................... |
| E | 3111 | hCoV-19/Japan/PG-0280/2020\|EPI_ISL_479960\|2020-03- | .................... | .................... | .................... | .................... | .................... | .................... |
| E | 3112 | hCoV-19/Japan/PG-0281/2020\|EPI_ISL_479961\|2020-03- | .................... | .................... | .................... | .................... | .................... | .................... |
| E | 3113 | hCoV-19/Japan/PG-0283/2020\|EPI_ISL_479962\|2020-03- | .................... | .................... | .................... | .................... | .................... | .................... |
| E | 3114 | hCoV-19/Japan/PG-0284/2020\|EPI_ISL_479963\|2020-03- | .................... | .................... | .................... | .................... | .................... | .................... |
| E | 3115 | hCoV-19/Japan/PG-0285/2020\|EPI_ISL_479964\|2020-03- | .................... | .................... | .................... | .................... | .................... | .................... |
| E | 3116 | hCoV-19/Japan/PG-0286/2020\|EPI_ISL_479965\|2020-03- | .................... | .................... | .................... | .................... | .................... | .................... |
| E | 3117 | hCoV-19/Japan/PG-0287/2020\|EPI_ISL_479966\|2020-03 | .................... | .................... | .................... | .................... | .................... | .................... |
| E | 3118 | hCoV-19/Japan/PG-0289/2020\|EPI_ISL_479967\|2020-03- | .................... | .................... | .................... | .................... | .................... | .................... |
| E | 3119 | hCoV-19/Japan/PG-0290/2020\|EPI_ISL_479968\|2020-03- | .................... | .................... | .................... | .................... | .................... | .................... |
| E | 3120 | hCoV-19/Japan/PG-0291/2020\|EPI_ISL_479969\|2020-03- | .................... | .................... | .................... | .................... | .................... | .................... |
| E | 3121 | hCoV-19/Japan/PG-0292/2020\|EPI_ISL_479970\|2020-03- | .................... | .................... | .................... | .................... | .................... | .................... |
| E | 3122 | hCoV-19/Japan/PG-0294/2020\|EPI_ISL_479971\|2020-03- | .................... | .................... | .................... | .................... | .................... | .................... |
| E | 3123 | hCoV-19/Japan/PG-0295/2020\|EPI_ISL_479972\|2020-03- | .................... | .................... | .................... | .................... | .................... | .................... |
| E | 3124 | hCoV-19/Japan/PG-0296/2020\|EPI_ISL_479973\|2020-03- | .................... | .................... | .................... | .................... | .................... | .................... |
| E | 3125 | hCoV-19/Japan/PG-0297/2020\|EPI_ISL_479974\|2020-03- | .................... | .................... | .................... | .................... | .................... | .................... |
| E | 3126 | hCoV-19/Japan/PG-0299/2020\|EPI_ISL_479975\|2020-03- | .................... | .................... | .................... | .................... | .................... | .................... |
| E | 3127 | hCoV-19/Japan/PG-0300/2020\|EPI_ISL_479976\|2020-03- | .................... | .................... | .................... | .................... | .................... | .................... |
| E | 3128 | hCoV-19/Japan/PG-0301/2020\|EPI_ISL_479977\|2020-03- | .................... | .................... | .................... | .................... | .................... | .................... |
| E | 3129 | hCoV-19/Japan/PG-0302/2020\|EPI_ISL_479978\|2020-03- | .................... | .................... | .................... | .................... | .................... | .................... |
| E | 3130 | hCoV-19/Japan/PG-0303/2020\|EPI_ISL_479979\|2020-03- | .................... | .................... | .................... | .................... | .................... | .................... |
| E | 3131 | hCoV-19/Japan/PG-0304/2020\|EPI_ISL_479980\|2020-03- | .................... | .................... | .................... | .................... | .................... | .................... |
| E | 3132 | hCoV-19/Japan/PG-0306/2020\|EPI_ISL_479981\|2020-03- | .................... | .................... | .................... | .................... | .................... | .................... |
| E | 3133 | hCoV-19/Japan/PG-0307/2020\|EPI_ISL_479982\|2020-03- | .................... | .................... | .................... | .................... | .................... | .................... |
| E | 3134 | hCoV-19/Japan/PG-0308/2020\|EPI_ISL_479983\|2020-03- | .................... | .................... | .................... | .................... | .................... | .................... |
| E | 3135 | hCoV-19/Japan/PG-0309/2020\|EPI_ISL_479984\|2020-03- | .................... | .................... | .................... | .................... | .................... | .................... |
| E | 3136 | hCoV-19/Japan/PG-0310/2020\|EPI_ISL_479985\|2020-03- | .................... | .................... | .................... | .................... | .................... | .................... |
| E | 3137 | hCoV-19/Japan/PG-0311/2020\|EPI_ISL_479986\|2020-03- | .................... | .................... | .................... | .................... | .................... | .................... |
| E | 3138 | hCoV-19/USA/FL-BPHL-0249/2020\|EPI_ISL_480900\|2020- | .................... | .................... | .................... | .................... | .................... | .................... |
| E | 3139 | hCoV-19/USA/FL-BPHL-0250/2020\|EPI_ISL_480901\|2020- | .................... | .................... | .................... | .................... | .................... | .................... |
| E | 3140 | hCoV-19/USA/FL-BPHL-0252/2020\|EPI_ISL_480903\|2020- | .................... | .................... | .................... | .................... | .................... | .................... |
| E | 3141 | hCoV-19/USA/FL-BPHL-0253/2020\|EPI_ISL_480904\|2020- | .................... | .................... | .................... | .................... | .................... | .................... |
| E | 3142 | hCoV-19/USA/FL-BPHL-0254/2020\|EPI_ISL_480905\|2020- | .................... | .................... | .................... | .................... | .................... | .................... |
| E | 3143 | hCoV-19/USA/FL-BPHL-0255/2020\|EPI_ISL_480906\|2020- | .................... | .................... | .................... | .................... | .................... | .................... |
| E | 3144 | hCoV-19/USA/FL-BPHL-0256/2020\|EPI_ISL_480907\|2020- | .................... | .................... | .................... | .................... | .................... | .................... |
| E | 3145 | hCoV-19/USA/FL-BPHL-0257/2020\|EPI_ISL_480908\|2020- | .................... | .................... | .................... | .................... | .................... | .................... |
| E | 3146 | hCoV-19/USA/FL-BPHL-0258/2020\|EPI_ISL_480909\|2020- | .................... | .................... | .................... | .................... | .................... | .................... |
| E | 3147 | hCoV-19/USA/FL-BPHL-0282/2020\|EPI_ISL_480933\|2020- | .................... | .................... | .................... | .................... | .................... | .................... |
| E | 3148 | hCoV-19/USA/FL-BPHL-0283/2020\|EPI_ISL_480934\|2020- | .................... | .................... | .................... | .................... | .................... | .................... |
| E | 3149 | hCoV-19/USA/FL-BPHL-0284/2020\|EPI_ISL_480935\|2020- | .................... | .................... | .................... | .................... | .................... | .................... |
| E | 3150 | hCoV-19/USA/FL-BPHL-0291/2020\|EPI_ISL_480942\|2020- | .................... | .................... | .................... | .................... | .................... | .................... |
| E | 3151 | hCoV-19/USA/FL-BPHL-0293/2020\|EPI_ISL_480944\|2020- | .................... | .................... | .................... | .................... | .................... | .................... |

| | | | | | | | | |
|---|---|---|---|---|---|---|---|---|
| E | 3152 hCoV-19/USA/FL-BPHL-0294/2020\|EPI_ISL_480945\|2020- | ···················· | ····T··············· | ···················· | ···················· | ···················· | ···················· | |
| E | 3153 hCoV-19/USA/FL-BPHL-0296/2020\|EPI_ISL_480947\|2020- | ···················· | ···················· | ···················· | ···················· | ···················· | ···················· | |
| E | 3154 hCoV-19/USA/FL-BPHL-0297/2020\|EPI_ISL_480948\|2020- | ···················· | ···················· | ···················· | ···················· | ···················· | ···················· | |
| E | 3155 hCoV-19/USA/FL-BPHL-0298/2020\|EPI_ISL_480949\|2020- | ···················· | ···················· | ···················· | ···················· | ···················· | ···················· | |
| E | 3156 hCoV-19/USA/FL-BPHL-0299/2020\|EPI_ISL_480950\|2020- | ···················· | ····T··············· | ···················· | ···················· | ···················· | ···················· | |
| E | 3157 hCoV-19/USA/FL-BPHL-0300/2020\|EPI_ISL_480951\|2020- | ···················· | ···················· | ···················· | ···················· | ···················· | ···················· | |
| E | 3165 hCoV-19/Spain/PV-IBV-003214/2020\|EPI_ISL_480959\|20 | ···················· | ···················· | ···················· | ···················· | ···················· | ···················· | |
| E | 3182 hCoV-19/USA/CA-ALSR-0476-SAN/2020\|EPI_ISL_483166\|2 | ···················· | ···················· | ···················· | ···················· | ···················· | ···················· | |
| E | 3183 hCoV-19/USA/CA-ALSR-0477-SAN/2020\|EPI_ISL_483167\|2 | ···················· | ···················· | ···················· | ···················· | ···················· | ···················· | |
| E | 3184 hCoV-19/USA/CA-ALSR-0479-SAN/2020\|EPI_ISL_483168\|2 | ···················· | ···················· | ···················· | ···················· | ···················· | ···················· | |
| E | 3185 hCoV-19/USA/CA-ALSR-0481-SAN/2020\|EPI_ISL_483169\|2 | ···················· | ···················· | ···················· | ···················· | ···················· | ···················· | |
| E | 3186 hCoV-19/USA/CA-ALSR-0483-SAN/2020\|EPI_ISL_483171\|2 | ···················· | ···················· | ···················· | ···················· | ···················· | ···················· | |
| E | 3187 hCoV-19/USA/CA-ALSR-0484-SAN/2020\|EPI_ISL_483172\|2 | ···················· | ···················· | ···················· | ···················· | ···················· | ···················· | |
| E | 3188 hCoV-19/USA/CA-ALSR-0489-SAN/2020\|EPI_ISL_483175\|2 | ···················· | ·A·················· | ···················· | ···················· | ···················· | ···················· | |
| E | 3189 hCoV-19/USA/CA-ALSR-0492-SAN/2020\|EPI_ISL_483177\|2 | ···················· | ···················· | ···················· | ···················· | ···················· | ···················· | |
| E | 3190 hCoV-19/USA/CA-ALSR-0493-SAN/2020\|EPI_ISL_483178\|2 | ···················· | ·A·················· | ···················· | ···················· | ···················· | ···················· | |
| E | 3191 hCoV-19/USA/CA-ALSR-0494-SAN/2020\|EPI_ISL_483179\|2 | ···················· | ···················· | ···················· | ···················· | ···················· | ···················· | |
| E | 3192 hCoV-19/USA/CA-ALSR-0495-IPL/2020\|EPI_ISL_483180\|2 | ···················· | ···················· | ···················· | ···················· | ···················· | ···················· | |
| E | 3193 hCoV-19/USA/CA-ALSR-0496-IPL/2020\|EPI_ISL_483181\|2 | ···················· | ···················· | ···················· | ···················· | ···················· | ···················· | |
| E | 3194 hCoV-19/USA/CA-ALSR-0497-IPL/2020\|EPI_ISL_483182\|2 | ···················· | ·A·················· | ···················· | ···················· | ···················· | ···················· | |
| E | 3195 hCoV-19/USA/CA-ALSR-0499-SAN/2020\|EPI_ISL_483183\|2 | ···················· | ···················· | ···················· | ···················· | ···················· | ···················· | |
| E | 3196 hCoV-19/USA/CA-ALSR-0500-IPL/2020\|EPI_ISL_483184\|2 | ···················· | ···················· | ···················· | ···················· | ···················· | ···················· | |
| E | 3197 hCoV-19/USA/CA-ALSR-0501-IPL/2020\|EPI_ISL_483185\|2 | ···················· | ···················· | ···················· | ···················· | ···················· | ···················· | |
| E | 3198 hCoV-19/USA/CA-ALSR-0502-SAN/2020\|EPI_ISL_483186\|2 | ···················· | ···················· | ···················· | ···················· | ···················· | ···················· | |
| E | 3199 hCoV-19/USA/CA-ALSR-0503-SAN/2020\|EPI_ISL_483187\|2 | ···················· | ···················· | ···················· | ···················· | ···················· | ···················· | |
| E | 3200 hCoV-19/Scotland/EDB5068/2020\|EPI_ISL_488000\|2020- | ···················· | ···················· | ···················· | ···················· | ···················· | ···················· | |
| E | 3201 hCoV-19/Scotland/EDB5516/2020\|EPI_ISL_488001\|2020- | ···················· | ···················· | ···················· | ···················· | ···················· | ···················· | |
| E | 3202 hCoV-19/Scotland/EDB4127/2020\|EPI_ISL_488002\|2020- | ···················· | ···················A | ···················· | ···················· | ···················· | ···················· | 1 |
| E | 3203 hCoV-19/England/NORT-2985C0/2020\|EPI_ISL_488003\|20 | ···················· | ···················· | ···················· | ···················· | ···················· | ···················· | |
| E | 3204 hCoV-19/England/NORT-29851B/2020\|EPI_ISL_488004\|20 | ···················· | ···················· | ···················· | ···················· | ···················· | ···················· | |
| E | 3205 hCoV-19/England/NORT-296B57/2020\|EPI_ISL_488005\|20 | ···················· | ···················· | ···················· | ···················· | ···················· | ···················· | |
| E | 3206 hCoV-19/England/NORT-297567/2020\|EPI_ISL_488007\|20 | ···················· | ···················· | ···················· | ···················· | ···················· | ···················· | |
| E | 3207 hCoV-19/England/NORT-29760A/2020\|EPI_ISL_488008\|20 | ···················· | ···················· | ···················· | ···················· | ···················· | ···················· | |
| E | 3208 hCoV-19/England/NORT-2973B8/2020\|EPI_ISL_488009\|20 | ···················· | ···················· | ···················· | ···················· | ···················· | ···················· | |
| E | 3209 hCoV-19/England/NORT-297479/2020\|EPI_ISL_488011\|20 | ···················· | ···················· | ···················· | ···················· | ···················· | ···················· | |
| E | 3210 hCoV-19/England/NORT-2984E1/2020\|EPI_ISL_488012\|20 | ···················· | ···················· | ···················· | ···················· | ···················· | ···················· | |
| E | 3211 hCoV-19/England/NORT-2985EE/2020\|EPI_ISL_488013\|20 | ···················· | ···················· | ···················· | ···················· | ···················· | ···················· | |
| E | 3212 hCoV-19/England/NORT-2969E4/2020\|EPI_ISL_488014\|20 | ···················· | ···················· | ···················· | ···················· | ···················· | ···················· | |
| E | 3213 hCoV-19/Northern Ireland/NIRE-101B76/2020\|EPI_ISL_ | ···················· | ···················· | ···················· | ···················· | ···················· | ···················· | |
| E | 3214 hCoV-19/Northern Ireland/NIRE-FD4A0/2020\|EPI_ISL_4 | ···················· | ···················· | ···················· | ···················· | ···················· | ···················· | |
| E | 3215 hCoV-19/Northern Ireland/NIRE-FFEFB/2020\|EPI_ISL_4 | ···················· | ···················· | ···················· | ···················· | ···················· | ···················· | |
| E | 3216 hCoV-19/Northern Ireland/NIRE-1034A6/2020\|EPI_ISL_ | ···················· | ···················· | ···················· | ···················· | ···················· | ···················· | |
| E | 3217 hCoV-19/Northern Ireland/NIRE-100324/2020\|EPI_ISL_ | ···················· | ···················· | ···················· | ···················· | ···················· | ···················· | |
| E | 3218 hCoV-19/Northern Ireland/NIRE-1000B4/2020\|EPI_ISL_ | ···················· | ···················· | ···················· | ···················· | ···················· | ···················· | |
| E | 3219 hCoV-19/Northern Ireland/NIRE-FFF43/2020\|EPI_ISL_4 | ···················· | ···················· | ···················· | ···················· | ···················· | ···················· | |
| E | 3220 hCoV-19/Northern Ireland/NIRE-101806/2020\|EPI_ISL_ | ···················· | ···················· | ···················· | ···················· | ···················· | ···················· | |
| E | 3221 hCoV-19/Northern Ireland/NIRE-104584/2020\|EPI_ISL_ | ···················· | ···················· | ···················· | ···················· | ···················· | ···················· | |
| E | 3222 hCoV-19/Northern Ireland/NIRE-FCEFE/2020\|EPI_ISL_4 | ···················· | ···················· | ···················· | ···················· | ···················· | ···················· | |
| E | 3223 hCoV-19/Northern Ireland/NIRE-102F5B/2020\|EPI_ISL_ | ···················· | ···················· | ···················· | ···················· | ···················· | ···················· | |
| E | 3224 hCoV-19/Northern Ireland/NIRE-FCD1F/2020\|EPI_ISL_4 | ···················· | ···················· | ···················· | ···················· | ···················· | ···················· | |
| E | 3225 hCoV-19/Northern Ireland/NIRE-FD2E2/2020\|EPI_ISL_4 | ···················· | ···················· | ···················· | ···················· | ···················· | ···················· | |
| E | 3226 hCoV-19/Northern Ireland/NIRE-100193/2020\|EPI_ISL_ | ···················· | ···················· | ···················· | ···················· | ···················· | ···················· | |
| E | 3227 hCoV-19/Northern Ireland/NIRE-1036CE/2020\|EPI_ISL_ | ···················· | ···················· | ···················· | ···················· | ···················· | ···················· | |
| E | 3228 hCoV-19/Northern Ireland/NIRE-1035C1/2020\|EPI_ISL_ | ···················· | ···················· | ···················· | ···················· | ···················· | ···················· | |
| E | 3229 hCoV-19/Northern Ireland/NIRE-1002FA/2020\|EPI_ISL_ | ···················· | ···················· | ···················· | ···················· | ···················· | ···················· | |
| E | 3230 hCoV-19/Northern Ireland/NIRE-1035EF/2020\|EPI_ISL_ | ···················· | ···················· | ···················· | ···················· | ···················· | ···················· | |
| E | 3231 hCoV-19/Northern Ireland/NIRE-FD1E5/2020\|EPI_ISL_4 | ···················· | ···················· | ···················· | ···················· | ···················· | ···················· | |
| E | 3232 hCoV-19/Northern Ireland/NIRE-10343D/2020\|EPI_ISL_ | ···················· | ···················· | ···················· | ···················· | ···················· | ···················· | |
| E | 3233 hCoV-19/Peru/LIM-INS-016/2020\|EPI_ISL_490315\|2020- | ···················· | ···················· | ···················· | ···················· | ···················· | ···················· | |
| E | 3234 hCoV-19/Peru/LIM-INS-017/2020\|EPI_ISL_490316\|2020- | ···················· | ···················· | ···················· | ···················· | ···················· | ···················· | |
| E | 3235 hCoV-19/England/CAMB-1B5458/2020\|EPI_ISL_490327\|20 | ···················· | ···················· | ···················· | ···················· | ···················· | ···················· | |
| E | 3236 hCoV-19/England/CAMB-1B5476/2020\|EPI_ISL_490328\|20 | ···················· | ···················· | ···················· | ···················· | ···················· | ···················· | |
| E | 3237 hCoV-19/England/CAMB-1B54A3/2020\|EPI_ISL_490329\|20 | ···················· | ···················· | ···················· | ···················· | ···················· | ···················· | |
| E | 3238 hCoV-19/Scotland/EDB1373/2020\|EPI_ISL_491600\|2020- | ···················· | ···················· | ···················· | ···················· | ···················· | ···················· | |
| E | 3239 hCoV-19/Scotland/EDB931/2020\|EPI_ISL_491602\|2020-0 | ···················· | ···················· | ···················· | ···················· | ···················· | ···················· | |
| E | 3240 hCoV-19/Scotland/EDB2360/2020\|EPI_ISL_491603\|2020- | ···················· | ·A·················· | ···················· | ···················· | ···················· | ···················· | |
| E | 3241 hCoV-19/Scotland/EDB869/2020\|EPI_ISL_491604\|2020-0 | ···················· | ·A·················· | ···················· | ···················· | ···················· | ···················· | |
| E | 3242 hCoV-19/Scotland/EDB510/2020\|EPI_ISL_491605\|2020-0 | ···················· | ···················· | ···················· | ···················· | ···················· | ···················· | |
| E | 3243 hCoV-19/Scotland/EDB1493/2020\|EPI_ISL_491606\|2020- | ···················· | ···················· | ···················· | ···················· | ···················· | ···················· | |
| E | 3244 hCoV-19/Scotland/EDB6445/2020\|EPI_ISL_491609\|2020- | ···················· | ···················· | ···················· | ···················· | ···················· | ···················· | |
| E | 3245 hCoV-19/Scotland/EDB1919/2020\|EPI_ISL_491610\|2020- | ···················· | ·A·················· | ···················· | ···················· | ···················· | ···················· | |
| E | 3246 hCoV-19/Scotland/EDB1898/2020\|EPI_ISL_491612\|2020- | ···················· | ···················A | ···················· | ···················· | ···················· | ···················· | 1 |
| E | 3247 hCoV-19/Scotland/EDB2354/2020\|EPI_ISL_491613\|2020- | ···················· | ·A·················· | ···················· | ···················· | ···················· | ···················· | |
| E | 3248 hCoV-19/Scotland/EDB3699/2020\|EPI_ISL_491614\|2020- | ···················· | ···················· | ···················· | ···················· | ···················· | ···················· | |
| E | 3249 hCoV-19/Scotland/EDB1143/2020\|EPI_ISL_491615\|2020- | ···················· | ···················· | ···················· | ···················· | ···················· | ···················· | |
| E | 3250 hCoV-19/Scotland/EDB6444/2020\|EPI_ISL_491617\|2020- | ···················· | ···················· | ···················· | ···················· | ···················· | ···················· | |
| E | 3251 hCoV-19/Scotland/EDB6740/2020\|EPI_ISL_491618\|2020- | ···················· | ···················· | ···················· | ···················· | ···················· | ···················· | |
| E | 3252 hCoV-19/Scotland/EDB961/2020\|EPI_ISL_491640\|2020-0 | ···················· | ·············A······ | ···················· | ···················· | ···················· | ···················· | |
| E | 3253 hCoV-19/Scotland/EDB1010/2020\|EPI_ISL_491643\|2020- | ···················· | ···················· | ···················· | ···················· | ···················· | ···················· | |
| E | 3254 hCoV-19/Scotland/EDB1371/2020\|EPI_ISL_491644\|2020- | ···················· | ···················· | ···················· | ···················· | ···················· | ···················· | |

| | | | | | | | | | |
|---|---|---|---|---|---|---|---|---|---|
| E | 3255 | hCoV-19/Scotland/EDB2286/2020\|EPI_ISL_491647\|2020- | .................... | .................... | .................... | .................... | .................... | .................... | |
| E | 3256 | hCoV-19/Scotland/EDB1353/2020\|EPI_ISL_491648\|2020- | .................... | .................... | .................... | .................... | .................... | .................... | |
| E | 3257 | hCoV-19/Scotland/EDB6742/2020\|EPI_ISL_491649\|2020- | .................... | ...................A | .................... | .................... | .................... | .................... | 1 |
| E | 3258 | hCoV-19/Scotland/EDB3824/2020\|EPI_ISL_491652\|2020- | .................... | .A.................. | .................... | .................... | .................... | .................... | |
| E | 3259 | hCoV-19/Scotland/EDB2009/2020\|EPI_ISL_491654\|2020- | .................... | .................... | .................... | .................... | .................... | .................... | |
| E | 3260 | hCoV-19/Scotland/EDB2061/2020\|EPI_ISL_491658\|2020- | .................... | .A.................. | .................... | .................... | .................... | .................... | |
| E | 3261 | hCoV-19/Scotland/EDB2549/2020\|EPI_ISL_491659\|2020- | .................... | .A.................. | .................... | .................... | .................... | .................... | |
| E | 3262 | hCoV-19/England/WSFT-25C10A6/2020\|EPI_ISL_492910\|2 | .................... | .................... | .................... | .................... | .................... | .................... | |
| E | 3263 | hCoV-19/England/WSFT-25C102E/2020\|EPI_ISL_492912\|2 | .................... | .................... | .................... | .................... | .................... | .................... | |
| E | 3264 | hCoV-19/England/WSFT-25C1343/2020\|EPI_ISL_492913\|2 | .................... | .................... | .................... | .................... | .................... | .................... | |
| E | 3265 | hCoV-19/England/LOND-124C610/2020\|EPI_ISL_492914\|2 | .................... | .................... | .................... | .................... | .................... | .................... | |
| E | 3266 | hCoV-19/England/WSFT-25C1334/2020\|EPI_ISL_492915\|2 | .................... | .................... | .................... | .................... | .................... | .................... | |
| E | 3267 | hCoV-19/England/NORT-28B0FC/2020\|EPI_ISL_492916\|20 | .................... | .................... | .................... | .................... | .................... | .................... | |
| E | 3268 | hCoV-19/England/NORT-28B092/2020\|EPI_ISL_492918\|20 | .................... | .................... | .................... | .................... | .................... | .................... | |
| E | 3269 | hCoV-19/England/NORT-28ACEA/2020\|EPI_ISL_492919\|20 | .................... | .................... | .................... | .................... | .................... | .................... | |
| E | 3270 | hCoV-19/England/NORT-28B047/2020\|EPI_ISL_492920\|20 | .................... | .................... | .................... | .................... | .................... | .................... | |
| E | 3271 | hCoV-19/England/NORT-28B065/2020\|EPI_ISL_492922\|20 | .................... | .................... | .................... | .................... | .................... | .................... | |
| E | 3272 | hCoV-19/England/NORT-28B28D/2020\|EPI_ISL_492923\|20 | .................... | .................... | .................... | .................... | .................... | .................... | |
| E | 3273 | hCoV-19/England/NORT-28C538/2020\|EPI_ISL_492924\|20 | .................... | .................... | .................... | .................... | .................... | .................... | |
| E | 3274 | hCoV-19/England/NORT-28AF5A/2020\|EPI_ISL_492925\|20 | .................... | .................... | .................... | .................... | .................... | .................... | |
| E | 3275 | hCoV-19/England/NORT-28C40E/2020\|EPI_ISL_492926\|20 | .................... | .................... | .................... | .................... | .................... | .................... | |
| E | 3276 | hCoV-19/England/NORT-28B1DB/2020\|EPI_ISL_492927\|20 | .................... | .................... | .................... | .................... | .................... | .................... | |
| E | 3277 | hCoV-19/England/NORT-28B1EA/2020\|EPI_ISL_492928\|20 | .................... | .................... | .................... | .................... | .................... | .................... | |
| E | 3278 | hCoV-19/England/NORT-28BFC2/2020\|EPI_ISL_492930\|20 | .................... | .................... | .................... | .................... | .................... | .................... | |
| E | 3279 | hCoV-19/England/NORT-28AEE4/2020\|EPI_ISL_492932\|20 | .................... | .................... | .................... | .................... | .................... | .................... | |
| E | 3280 | hCoV-19/England/NORT-28B056/2020\|EPI_ISL_492934\|20 | ..................Y. | .................... | .................... | .................... | .................... | .................... | 1 |
| E | 3281 | hCoV-19/England/NORT-28AF69/2020\|EPI_ISL_492935\|20 | .................... | .................... | .................... | .................... | .................... | .................... | |
| E | 3282 | hCoV-19/England/NORT-28B1BD/2020\|EPI_ISL_492936\|20 | .................... | .................... | .................... | .................... | .................... | .................... | |
| E | 3283 | hCoV-19/England/NORT-28B108/2020\|EPI_ISL_492937\|20 | .................... | .................... | .................... | .................... | .................... | .................... | |
| E | 3284 | hCoV-19/England/NORT-28ADAB/2020\|EPI_ISL_492938\|20 | .................... | ...................A | .................... | .................... | .................... | .................... | 1 |
| E | 3285 | hCoV-19/England/NORT-28B26F/2020\|EPI_ISL_492939\|20 | .................... | .................... | .................... | .................... | .................... | .................... | |
| E | 3286 | hCoV-19/England/NORT-28AD8D/2020\|EPI_ISL_492942\|20 | .................... | .................... | .................... | .................... | .................... | .................... | |
| E | 3287 | hCoV-19/England/NORT-28AE5D/2020\|EPI_ISL_492943\|20 | .................... | .................... | .................... | .................... | .................... | .................... | |
| E | 3288 | hCoV-19/England/NORT-28C055/2020\|EPI_ISL_492944\|20 | .................... | .................... | .................... | .................... | .................... | .................... | |
| E | 3289 | hCoV-19/England/NORT-28AE99/2020\|EPI_ISL_492945\|20 | .................... | .................... | .................... | .................... | .................... | .................... | |
| E | 3290 | hCoV-19/England/NORT-28ACF9/2020\|EPI_ISL_492946\|20 | .................... | .................... | .................... | .................... | .................... | .................... | |
| E | 3291 | hCoV-19/England/NORT-28ADD8/2020\|EPI_ISL_492947\|20 | .................... | .................... | .................... | .................... | .................... | .................... | |
| E | 3292 | hCoV-19/England/NORT-28AF4B/2020\|EPI_ISL_492948\|20 | .................... | .................... | .................... | .................... | .................... | .................... | |
| E | 3293 | hCoV-19/England/NORT-28C2D7/2020\|EPI_ISL_492949\|20 | .................... | .A.................. | .................... | .................... | .................... | .................... | |
| E | 3294 | hCoV-19/England/NORT-28AD05/2020\|EPI_ISL_492970\|20 | .................... | .................... | .................... | .................... | .................... | .................... | |
| E | 3295 | hCoV-19/England/NORT-28AEA8/2020\|EPI_ISL_492973\|20 | .................... | .................... | .................... | .................... | .................... | .................... | |
| E | 3296 | hCoV-19/England/NORT-28B083/2020\|EPI_ISL_492974\|20 | .................... | .................... | .................... | .................... | .................... | .................... | |
| E | 3297 | hCoV-19/England/NORT-28C028/2020\|EPI_ISL_492975\|20 | .................... | .................... | .................... | .................... | .................... | .................... | |
| E | 3298 | hCoV-19/England/NORT-28AD9C/2020\|EPI_ISL_492976\|20 | .................... | .................... | .................... | .................... | .................... | .................... | |
| E | 3299 | hCoV-19/England/NORT-28AE4E/2020\|EPI_ISL_492977\|20 | .................... | .................... | .................... | .................... | .................... | .................... | |
| E | 3300 | hCoV-19/Singapore/480/2020\|EPI_ISL_492978\|2020-03- | .................... | .................... | .................... | .................... | .................... | .................... | |
| E | 3301 | hCoV-19/Singapore/484/2020\|EPI_ISL_492979\|2020-03- | .................... | .................... | .................... | .................... | .................... | .................... | |
| E | 3302 | hCoV-19/Italy/VEN-UniVR-3/2020\|EPI_ISL_492982\|2020 | .................... | .................... | .................... | .................... | .................... | .................... | |
| E | 3303 | hCoV-19/Italy/VEN-UniVR-4/2020\|EPI_ISL_492983\|2020 | .................... | .................... | .................... | .................... | .................... | .................... | |
| E | 3304 | hCoV-19/Italy/VEN-UniVR-5/2020\|EPI_ISL_492984\|2020 | .................... | .................... | .................... | .................... | .................... | .................... | |
| E | 3305 | hCoV-19/Italy/VEN-UniVR-6/2020\|EPI_ISL_492985\|2020 | .................... | .................... | .................... | .................... | .................... | .................... | |
| E | 3306 | hCoV-19/Italy/VEN-UniVR-7/2020\|EPI_ISL_492986\|2020 | .................... | .................... | .................... | .................... | .................... | .................... | |
| E | 3307 | hCoV-19/Italy/VEN-UniVR-8/2020\|EPI_ISL_492987\|2020 | .................... | .................... | .................... | .................... | .................... | .................... | |
| E | 3308 | hCoV-19/Latvia/043/2020\|EPI_ISL_492988\|2020-06-30 | .................... | .................... | .................... | .................... | .................... | .................... | |
| E | 3309 | hCoV-19/Latvia/066/2020\|EPI_ISL_492989\|2020-07-08 | .................... | .................... | .................... | .................... | .................... | .................... | |
| E | 3310 | hCoV-19/USA/CA-IGI-0127/2020\|EPI_ISL_495149\|2020-0 | .................... | .................... | .................... | .................... | .................... | .................... | |
| E | 3311 | hCoV-19/USA/CA-IGI-0128/2020\|EPI_ISL_495150\|2020-0 | .................... | .................... | .................... | .................... | .................... | .................... | |
| E | 3312 | hCoV-19/India/TG-CCMB-L1011/2020\|EPI_ISL_495161\|20 | .................... | .................... | .................... | .................... | .................... | .................... | |
| E | 3313 | hCoV-19/India/TG-CCMB-L1023/2020\|EPI_ISL_495162\|20 | .................... | .................... | .................... | .................... | .................... | .................... | |
| E | 3314 | hCoV-19/India/TG-CCMB-L1657/2020\|EPI_ISL_495163\|20 | .................... | .................... | .................... | .................... | .................... | .................... | |
| E | 3315 | hCoV-19/India/TG-CCMB-L1822-ONT/2020\|EPI_ISL_49516 | .................... | .................... | .................... | .................... | .................... | .................... | |
| E | 3316 | hCoV-19/India/TG-CCMB-L1824-ONT/2020\|EPI_ISL_49516 | .................... | .................... | .................... | .................... | .................... | .................... | |
| E | 3317 | hCoV-19/India/TG-CCMB-L1864/2020\|EPI_ISL_495166\|20 | .................... | .................... | .................... | .................... | .................... | .................... | |
| E | 3318 | hCoV-19/India/TG-CCMB-M172/2020\|EPI_ISL_495189\|202 | .................... | .................... | .................... | .................... | .................... | .................... | |
| E | 3319 | hCoV-19/India/TG-CCMB-M175/2020\|EPI_ISL_495190\|202 | .................... | .................... | .................... | .................... | .................... | .................... | |
| E | 3320 | hCoV-19/India/TG-CCMB-M181/2020\|EPI_ISL_495191\|202 | .................... | .................... | .................... | .................... | .................... | .................... | |
| E | 3321 | hCoV-19/India/TG-CCMB-M2/2020\|EPI_ISL_495192\|2020- | .................... | .................... | .................... | .................... | .................... | .................... | |
| E | 3322 | hCoV-19/India/TG-CCMB-M200/2020\|EPI_ISL_495193\|202 | .................... | .................... | .................... | .................... | .................... | .................... | |
| E | 3323 | hCoV-19/India/TG-CCMB-M22/2020\|EPI_ISL_495194\|2020 | .................... | .................... | .................... | .................... | .................... | .................... | |
| E | 3324 | hCoV-19/India/TG-CCMB-M243/2020\|EPI_ISL_495195\|202 | .................... | .A.................. | .................... | .................... | .................... | ...A................ | |
| E | 3325 | hCoV-19/India/TG-CCMB-M25/2020\|EPI_ISL_495196\|2020 | .................... | .................... | .................... | .................... | .................... | .................... | |
| E | 3327 | hCoV-19/India/TG-CCMB-M343/2020\|EPI_ISL_495198\|202 | .................... | .................... | .................... | .................... | .................... | .................... | |
| E | 3328 | hCoV-19/India/TG-CCMB-M357/2020\|EPI_ISL_495199\|202 | .................... | .................... | .................... | .................... | .................... | .................... | |
| E | 3329 | hCoV-19/Japan/PG-0222/2020\|EPI_ISL_479921\|2020-03- | .................... | .................... | .................... | .................... | .................... | .................... | |
| E | 3330 | hCoV-19/Japan/PG-0223/2020\|EPI_ISL_479922\|2020-03- | .................... | .................... | .................... | .................... | .................... | .................... | |
| E | 3331 | hCoV-19/Japan/PG-0224/2020\|EPI_ISL_479923\|2020-03- | .................... | .................... | .................... | .................... | .................... | .................... | |
| E | 3332 | hCoV-19/Japan/PG-0227/2020\|EPI_ISL_479924\|2020-03- | .................... | .................... | .................... | .................... | .................... | .................... | |
| E | 3333 | hCoV-19/Japan/PG-0229/2020\|EPI_ISL_479925\|2020-03- | .................... | .................... | .................... | .................... | .................... | .................... | |
| E | 3334 | hCoV-19/Japan/PG-0230/2020\|EPI_ISL_479926\|2020-03- | .................... | .................... | .................... | .................... | .................... | .................... | |
| E | 3335 | hCoV-19/Japan/PG-0232/2020\|EPI_ISL_479927\|2020-03- | .................... | .................... | .................... | .................... | .................... | .................... | |

| | | | | | | | | |
|---|---|---|---|---|---|---|---|---|
| E | 3336 | hCoV-19/Japan/PG-0233/2020\|EPI_ISL_479928\|2020-03- | ···················· | ···················· | ···················· | ···················· | ···················· | ···················· |
| E | 3337 | hCoV-19/Japan/PG-0234/2020\|EPI_ISL_479929\|2020-03- | ···················· | ···················· | ···················· | ···················· | ···················· | ···················· |
| E | 3338 | hCoV-19/Japan/PG-0235/2020\|EPI_ISL_479930\|2020-03- | ···················· | ···················· | ···················· | ···················· | ···················· | ···················· |
| E | 3339 | hCoV-19/Japan/PG-0237/2020\|EPI_ISL_479931\|2020-03- | ···················· | ···················· | ···················· | ···················· | ···················· | ···················· |
| E | 3340 | hCoV-19/Japan/PG-0241/2020\|EPI_ISL_479932\|2020-03- | ···················· | ···················· | ···················· | ···················· | ···················· | ···················· |
| E | 3341 | hCoV-19/Japan/PG-0243/2020\|EPI_ISL_479933\|2020-03- | ···················· | ···················· | ···················· | ···················· | ···················· | ···················· |
| E | 3342 | hCoV-19/Japan/PG-0246/2020\|EPI_ISL_479934\|2020-03- | ···················· | ···················· | ···················· | ···················· | ···················· | ···················· |
| E | 3343 | hCoV-19/Japan/PG-0247/2020\|EPI_ISL_479935\|2020-03- | ···················· | ···················· | ···················· | ···················· | ···················· | ···················· |
| E | 3344 | hCoV-19/Japan/PG-0251/2020\|EPI_ISL_479936\|2020-03- | ···················· | ···················· | ···················· | ···················· | ···················· | ···················· |
| E | 3345 | hCoV-19/Japan/PG-0252/2020\|EPI_ISL_479937\|2020-03- | ···················· | ···················· | ···················· | ···················· | ···················· | ···················· |
| E | 3346 | hCoV-19/Japan/PG-0253/2020\|EPI_ISL_479938\|2020-03- | ···················· | ···················· | ···················· | ···················· | ···················· | ···················· |
| E | 3347 | hCoV-19/Japan/PG-0254/2020\|EPI_ISL_479939\|2020-03- | ···················· | ···················· | ···················· | ···················· | ···················· | ···················· |
| E | 3348 | hCoV-19/Japan/PG-0255/2020\|EPI_ISL_479940\|2020-03- | ···················· | ···················· | ···················· | ···················· | ···················· | ···················· |
| E | 3349 | hCoV-19/Japan/PG-0256/2020\|EPI_ISL_479941\|2020-03- | ···················· | ···················· | ···················· | ···················· | ···················· | ···················· |
| E | 3350 | hCoV-19/Japan/PG-0258/2020\|EPI_ISL_479942\|2020-03- | ···················· | ···················· | ···················· | ···················· | ···················· | ···················· |
| E | 3351 | hCoV-19/USA/FL-BPHL-0242/2020\|EPI_ISL_480893\|2020- | ···················· | ···················· | ···················· | ···················· | ···················· | ···················· |
| E | 3352 | hCoV-19/USA/FL-BPHL-0245/2020\|EPI_ISL_480896\|2020- | ···················· | ···················· | ···················· | ···················· | ···················· | ···················· |
| E | 3353 | hCoV-19/USA/FL-BPHL-0246/2020\|EPI_ISL_480897\|2020- | ···················· | ···················· | ···················· | ···················· | ···················· | ···················· |
| E | 3354 | hCoV-19/USA/FL-BPHL-0247/2020\|EPI_ISL_480898\|2020- | ···················· | ···················· | ···················· | ···················· | ···················· | ···················· |
| E | 3355 | hCoV-19/USA/FL-BPHL-0248/2020\|EPI_ISL_480899\|2020- | ···················· | ···················· | ···················· | ···················· | ···················· | ···················· |
| E | 3356 | hCoV-19/USA/FL-BPHL-0251/2020\|EPI_ISL_480902\|2020- | ···················· | ···················· | ···················· | ···················· | ···················· | ···················· |
| E | 3357 | hCoV-19/USA/FL-BPHL-0259/2020\|EPI_ISL_480910\|2020- | ···················· | ···················· | ···················· | ···················· | ···················· | ···················· |
| E | 3358 | hCoV-19/USA/FL-BPHL-0262/2020\|EPI_ISL_480913\|2020- | ···················· | ···················· | ···················· | ···················· | ···················· | ···················· |
| E | 3359 | hCoV-19/USA/FL-BPHL-0263/2020\|EPI_ISL_480914\|2020- | ···················· | ···················· | ···················· | ···················· | ···················· | ···················· |
| E | 3360 | hCoV-19/USA/FL-BPHL-0265/2020\|EPI_ISL_480916\|2020- | ···················· | ···················· | ···················· | ···················· | ···················· | ···················· |
| E | 3361 | hCoV-19/USA/FL-BPHL-0266/2020\|EPI_ISL_480917\|2020- | ···················· | ···················· | ···················· | ···················· | ···················· | ···················· |
| E | 3362 | hCoV-19/USA/FL-BPHL-0267/2020\|EPI_ISL_480918\|2020- | ···················· | ···················· | ···················· | ···················· | ···················· | ···················· |
| E | 3363 | hCoV-19/USA/FL-BPHL-0270/2020\|EPI_ISL_480921\|2020- | ···················· | ···················· | ···················· | ···················· | ···················· | ···················· |
| E | 3364 | hCoV-19/USA/FL-BPHL-0271/2020\|EPI_ISL_480922\|2020- | ···················· | ···················· | ···················· | ···················· | ···················· | ···················· |
| E | 3365 | hCoV-19/USA/FL-BPHL-0275/2020\|EPI_ISL_480926\|2020- | ···················· | ···················· | ···················· | ···················· | ···················· | ···················· |
| E | 3366 | hCoV-19/USA/FL-BPHL-0276/2020\|EPI_ISL_480927\|2020- | ···················· | ···················· | ···················· | ···················· | ···················· | ···················· |
| E | 3367 | hCoV-19/USA/FL-BPHL-0277/2020\|EPI_ISL_480928\|2020- | ···················· | ···················· | ···················· | ···················· | ···················· | ···················· |
| E | 3368 | hCoV-19/Australia/SAP321/2020\|EPI_ISL_483101\|2020- | ···················· | ···················· | ···················· | ···················· | ···················· | ···················· |
| E | 3369 | hCoV-19/Australia/SAP327/2020\|EPI_ISL_483105\|2020- | ···················· | ···················· | ···················· | ···················· | ···················· | ···················· |
| E | 3370 | hCoV-19/Australia/SAP330/2020\|EPI_ISL_483108\|2020- | ···················· | ···················· | ···················· | ···················· | ···················· | ···················· |
| E | 3371 | hCoV-19/Australia/SAP331/2020\|EPI_ISL_483109\|2020- | ···················· | ···················· | ···················· | ···················· | ···················· | ···················· |
| E | 3372 | hCoV-19/Australia/SAP334/2020\|EPI_ISL_483111\|2020- | ···················· | ···················· | ···················· | ···················· | ···················· | ···················· |
| E | 3373 | hCoV-19/Australia/SAP336/2020\|EPI_ISL_483113\|2020- | ···················· | ···················· | ···················· | ···················· | ···················· | ···················· |
| E | 3374 | hCoV-19/Australia/SAP339/2020\|EPI_ISL_483116\|2020- | ···················· | ···················· | ···················· | ···················· | ···················· | ···················· |
| E | 3375 | hCoV-19/Australia/SAP340/2020\|EPI_ISL_483117\|2020- | ···················· | ···················· | ···················· | ···················· | ···················· | ···················· |
| E | 3376 | hCoV-19/Australia/SAP342/2020\|EPI_ISL_483119\|2020- | ···················· | ···················· | ···················· | ···················· | ···················· | ···················· |
| E | 3377 | hCoV-19/Australia/SAP344/2020\|EPI_ISL_483120\|2020- | ···················· | ···················· | ···················· | ···················· | ···················· | ···················· |
| E | 3378 | hCoV-19/Australia/SAP345/2020\|EPI_ISL_483121\|2020- | ···················· | ···················· | ···················· | ···················· | ···················· | ···················· |
| E | 3379 | hCoV-19/Australia/SAP346/2020\|EPI_ISL_483122\|2020- | ···················· | ···················· | ···················· | ···················· | ···················· | ···················· |
| E | 3380 | hCoV-19/Australia/SAP349/2020\|EPI_ISL_483124\|2020- | ···················· | ···················· | ···················· | ···················· | ···················· | ···················· |
| E | 3381 | hCoV-19/Australia/SAP350/2020\|EPI_ISL_483125\|2020- | ···················· | ···················· | ···················· | ···················· | ···················· | ···················· |
| E | 3382 | hCoV-19/Australia/SAP352/2020\|EPI_ISL_483126\|2020- | ···················· | ···················· | ···················· | ···················· | ···················· | ···················· |
| E | 3383 | hCoV-19/Australia/SAP354/2020\|EPI_ISL_483127\|2020- | ···················· | ···················· | ···················· | ···················· | ···················· | ···················· |
| E | 3384 | hCoV-19/Australia/SAP357/2020\|EPI_ISL_483129\|2020- | ···················· | ···················· | ···················· | ···················· | ···················· | ···················· |
| E | 3385 | hCoV-19/Australia/SAP360/2020\|EPI_ISL_483132\|2020- | ···················· | ···················· | ···················· | ···················· | ···················· | ···················· |
| E | 3386 | hCoV-19/Australia/SAP366/2020\|EPI_ISL_483137\|2020- | ···················· | ···················· | ···················· | ···················· | ···················· | ···················· |
| E | 3387 | hCoV-19/USA/CA-ALSR-0469-SAN/2020\|EPI_ISL_483160\|2 | ···················· | ···················· | ···················· | ···················· | ···················· | ···················· |
| E | 3388 | hCoV-19/USA/CA-ALSR-0470-SAN/2020\|EPI_ISL_483161\|2 | ···················· | ···················· | ···················· | ···················· | ···················· | ···················· |
| E | 3389 | hCoV-19/USA/CA-ALSR-0472-SAN/2020\|EPI_ISL_483162\|2 | ···················· | ·A·················· | ···················· | ···················· | ···················· | ···················· |
| E | 3390 | hCoV-19/USA/CA-ALSR-0473-SAN/2020\|EPI_ISL_483163\|2 | ···················· | ·A·················· | ···················· | ···················· | ···················· | ···················· |
| E | 3391 | hCoV-19/USA/CA-ALSR-0474-SAN/2020\|EPI_ISL_483164\|2 | ···················· | ·A·················· | ···················· | ···················· | ···················· | ···················· |
| E | 3392 | hCoV-19/USA/CA-ALSR-0475-SAN/2020\|EPI_ISL_483165\|2 | ···················· | ···················· | ···················· | ···················· | ···················· | ···················· |
| E | 3393 | hCoV-19/USA/CA-ALSR-0505-SAN/2020\|EPI_ISL_483188\|2 | ···················· | ·A·················· | ···················· | ···················· | ···················· | ···················· |
| E | 3394 | hCoV-19/USA/CA-ALSR-0506-SAN/2020\|EPI_ISL_483189\|2 | ···················· | ···················· | ···················· | ···················· | ···················· | ···················· |
| E | 3395 | hCoV-19/USA/CA-ALSR-0508-SAN/2020\|EPI_ISL_483190\|2 | ···················· | ···················· | ···················· | ···················· | ···················· | ···················· |
| E | 3396 | hCoV-19/USA/CA-ALSR-0512-SAN/2020\|EPI_ISL_483191\|2 | ···················· | ···················· | ···················· | ···················· | ···················· | ···················· |
| E | 3397 | hCoV-19/USA/CA-ALSR-0513-SAN/2020\|EPI_ISL_483192\|2 | ···················· | ···················· | ···················· | ···················· | ···················· | ···················· |
| E | 3398 | hCoV-19/USA/CA-ALSR-0515-SAN/2020\|EPI_ISL_483194\|2 | ···················· | ···················· | ···················· | ···················· | ···················· | ···················· |
| E | 3399 | hCoV-19/USA/CA-ALSR-0516-SAN/2020\|EPI_ISL_483195\|2 | ···················· | ·A·················· | ···················· | ···················· | ···················· | ···················· |
| E | 3400 | hCoV-19/USA/CA-ALSR-0518-SAN/2020\|EPI_ISL_483197\|2 | ···················· | ···················· | ···················· | ···················· | ···················· | ···················· |
| E | 3401 | hCoV-19/USA/CA-ALSR-0519-IPL/2020\|EPI_ISL_483198\|2 | ···················· | ···················· | ···················· | ···················· | ···················· | ···················· |
| E | 3402 | hCoV-19/USA/CA-ALSR-0521-IPL/2020\|EPI_ISL_483199\|2 | ···················· | ···················· | ···················· | ···················· | ···················· | ···················· |
| E | 3403 | hCoV-19/England/NOTT-1125CF/2020\|EPI_ISL_484400\|20 | ···················· | ···················· | ···················· | ···················· | ···················· | ···················· |
| E | 3404 | hCoV-19/England/NOTT-1125DE/2020\|EPI_ISL_484401\|20 | ···················· | ···················· | ···················· | ···················· | ···················· | ···················· |
| E | 3405 | hCoV-19/England/NOTT-1125ED/2020\|EPI_ISL_484402\|20 | ···················· | ···················· | ···················· | ···················· | ···················· | ···················· |
| E | 3406 | hCoV-19/England/NOTT-112644/2020\|EPI_ISL_484406\|20 | ···················· | ···················· | ···················· | ···················· | ···················· | ···················· |
| E | 3407 | hCoV-19/England/PORT-2D115E/2020\|EPI_ISL_484407\|20 | ···················· | ···················· | ···················· | ···················· | ···················· | ···················· |
| E | 3408 | hCoV-19/England/PORT-2D158F/2020\|EPI_ISL_484410\|20 | ···················· | ···················· | ···················· | ···················· | ···················· | ···················· |
| E | 3409 | hCoV-19/England/PORT-2D15AD/2020\|EPI_ISL_484411\|20 | ···················· | ···················· | ···················· | ···················· | ···················· | ···················· |
| E | 3410 | hCoV-19/England/PORT-2D15DA/2020\|EPI_ISL_484412\|20 | ···················· | ···················· | ···················· | ···················· | ···················· | ···················· |
| E | 3411 | hCoV-19/England/PORT-2D15E9/2020\|EPI_ISL_484413\|20 | ···················· | ···················· | ···················· | ···················· | ···················· | ···················· |
| E | 3412 | hCoV-19/England/PORT-2D1613/2020\|EPI_ISL_484415\|20 | ···················· | ···················· | ···················· | ···················· | ···················· | ···················· |
| E | 3413 | hCoV-19/England/PORT-2D1631/2020\|EPI_ISL_484416\|20 | ···················· | ···················· | ···················· | ···················· | ···················· | ···················· |
| E | 3414 | hCoV-19/England/PORT-2D1640/2020\|EPI_ISL_484417\|20 | ···················· | ···················· | ···················· | ···················· | ···················· | ···················· |
| E | 3415 | hCoV-19/England/PORT-2D167D/2020\|EPI_ISL_484418\|20 | ···················· | ···················· | ···················· | ···················· | ···················· | ···················· |

| | | | | | | | | |
|---|---|---|---|---|---|---|---|---|
| E | 3416 | hCoV-19/England/PORT-2D17C5/2020\|EPI_ISL_484419\|20 | ···················· | ···················· | ···················· | ···················· | ···················· | ···················· |
| E | 3417 | hCoV-19/England/PORT-2D180E/2020\|EPI_ISL_484421\|20 | ···················· | ···················· | ···················· | ···················· | ···················· | ···················· |
| E | 3418 | hCoV-19/England/PORT-2D181D/2020\|EPI_ISL_484422\|20 | ···················· | ···················· | ···················· | ···················· | ···················· | ···················· |
| E | 3419 | hCoV-19/England/PORT-2D183B/2020\|EPI_ISL_484423\|20 | ···················· | ···················· | ···················· | ···················· | ···················· | ···················· |
| E | 3420 | hCoV-19/England/PORT-2D184A/2020\|EPI_ISL_484424\|20 | ···················· | ···················· | ···················· | ···················· | ···················· | ···················· |
| E | 3421 | hCoV-19/England/PORT-2D1877/2020\|EPI_ISL_484427\|20 | ···················· | ···················· | ···················· | ···················· | ···················· | ···················· |
| E | 3422 | hCoV-19/England/PORT-2D1886/2020\|EPI_ISL_484428\|20 | ···················· | ···················· | ···················· | ···················· | ···················· | ···················· |
| E | 3423 | hCoV-19/England/PORT-2D18A4/2020\|EPI_ISL_484429\|20 | ···················· | ···················· | ···················· | ···················· | ···················· | ···················· |
| E | 3424 | hCoV-19/England/PORT-2D18B3/2020\|EPI_ISL_484430\|20 | ···················· | ···················· | ···················· | ···················· | ···················· | ···················· |
| E | 3425 | hCoV-19/England/SHEF-C1038/2020\|EPI_ISL_484435\|202 | ···················· | ···················· | ···················· | ···················· | ···················· | ···················· |
| E | 3426 | hCoV-19/England/SHEF-C3E2D/2020\|EPI_ISL_484446\|202 | ···················· | ···················· | ···················· | ···················· | ···················· | ···················· |
| E | 3427 | hCoV-19/England/SHEF-C427B/2020\|EPI_ISL_484449\|202 | ···················· | ···················· | ···················· | ···················· | ···················· | ···················· |
| E | 3428 | hCoV-19/England/SHEF-C44FD/2020\|EPI_ISL_484451\|202 | ···················· | ···················· | ···················· | ···················· | ···················· | ···················· |
| E | 3429 | hCoV-19/England/SHEF-C4D98/2020\|EPI_ISL_484452\|202 | ···················· | ···················· | ···················· | ···················· | ···················· | ···················· |
| E | 3430 | hCoV-19/England/SHEF-C54DE/2020\|EPI_ISL_484453\|202 | ···················· | ···················· | ···················· | ···················· | ···················· | ···················· |
| E | 3431 | hCoV-19/England/SHEF-C568D/2020\|EPI_ISL_484454\|202 | ···················· | ···················· | ···················· | ···················· | ···················· | ···················· |
| E | 3432 | hCoV-19/England/SHEF-C578A/2020\|EPI_ISL_484456\|202 | ···················· | ···················· | ···················· | ···················· | ···················· | ···················· |
| E | 3433 | hCoV-19/England/SHEF-C5B9D/2020\|EPI_ISL_484459\|202 | ···················· | ···················· | ···················· | ···················· | ···················· | ···················· |
| E | 3434 | hCoV-19/England/SHEF-C5D1F/2020\|EPI_ISL_484460\|202 | ···················· | ···················· | ···················· | ···················· | ···················· | ···················· |
| E | 3435 | hCoV-19/England/SHEF-C61E5/2020\|EPI_ISL_484461\|202 | ···················· | ···················· | ···················· | ···················· | ···················· | ···················· |
| E | 3436 | hCoV-19/England/SHEF-C623D/2020\|EPI_ISL_484462\|202 | ···················· | ···················· | ···················· | ···················· | ···················· | ···················· |
| E | 3437 | hCoV-19/England/SHEF-C7630/2020\|EPI_ISL_484469\|202 | ···················· | ···················· | ···················· | ···················· | ···················· | ···················· |
| E | 3438 | hCoV-19/England/SHEF-C7876/2020\|EPI_ISL_484470\|202 | ···················· | ···················· | ···················· | ···················· | ···················· | ···················· |
| E | 3439 | hCoV-19/England/SHEF-C84F9/2020\|EPI_ISL_484473\|202 | ···················· | ···················· | ···················· | ···················· | ···················· | ···················· |
| E | 3440 | hCoV-19/England/SHEF-C8945/2020\|EPI_ISL_484474\|202 | ···················· | ···················· | ···················· | ···················· | ···················· | ···················· |
| E | 3441 | hCoV-19/England/SHEF-C8A8E/2020\|EPI_ISL_484475\|202 | ···················· | ···················· | ···················· | ···················· | ···················· | ···················· |
| E | 3442 | hCoV-19/England/SHEF-C8D1C/2020\|EPI_ISL_484476\|202 | ···················· | ···················· | ···················· | ···················· | ···················· | ···················· |
| E | 3443 | hCoV-19/England/SHEF-C913D/2020\|EPI_ISL_484478\|202 | ···················· | ···················· | ···················· | ···················· | ···················· | ···················· |
| E | 3444 | hCoV-19/England/SHEF-C9847/2020\|EPI_ISL_484482\|202 | ···················· | ···················· | ···················· | ···················· | ···················· | ···················· |
| E | 3445 | hCoV-19/England/SHEF-C9A8D/2020\|EPI_ISL_484484\|202 | ···················· | ···················· | ···················· | ···················· | ···················· | ···················· |
| E | 3446 | hCoV-19/England/SHEF-CA970/2020\|EPI_ISL_484487\|202 | ···················· | ···················· | ···················· | ···················· | ···················· | ···················· |
| E | 3447 | hCoV-19/England/SHEF-CAB7A/2020\|EPI_ISL_484489\|202 | ···················· | ···················· | ···················· | ···················· | ···················· | ···················· |
| E | 3448 | hCoV-19/England/SHEF-CB53F/2020\|EPI_ISL_484494\|202 | ···················· | ···················· | ···················· | ···················· | ···················· | ···················· |
| E | 3449 | hCoV-19/England/SHEF-CC255/2020\|EPI_ISL_484495\|202 | ···················· | ···················· | ···················· | ···················· | ···················· | ···················· |
| E | 3450 | hCoV-19/Senegal/66298/2020\|EPI_ISL_485708\|2020-06- | ···················· | ···················· | ···················· | ···················· | ···················· | ···················· |
| E | 3451 | hCoV-19/Senegal/66458/2020\|EPI_ISL_485710\|2020-06- | ···················· | ···················· | ···················· | ···················· | ···················· | ···················· |
| E | 3452 | hCoV-19/Senegal/65428/2020\|EPI_ISL_485712\|2020-06- | ···················· | ···················· | ···················· | ···················· | ···················· | ···················· |
| E | 3453 | hCoV-19/Senegal/65601/2020\|EPI_ISL_485713\|2020-06- | ···················· | ···················· | ···················· | ···················· | ···················· | ···················· |
| E | 3454 | hCoV-19/Senegal/64033/2020\|EPI_ISL_485715\|2020-06- | ···················· | ···················· | ···················· | ···················· | ···················· | ···················· |
| E | 3455 | hCoV-19/Senegal/64076/2020\|EPI_ISL_485716\|2020-06- | ···················· | ···················· | ···················· | ···················· | ···················· | ···················· |
| E | 3456 | hCoV-19/Senegal/64096/2020\|EPI_ISL_485717\|2020-06- | ···················· | ···················· | ···················· | ···················· | ···················· | ···················· |
| E | 3457 | hCoV-19/Northern Ireland/NIRE-1038B9/2020\|EPI_ISL_ | ···················· | ···················· | ···················· | ···················· | ···················· | ···················· |
| E | 3458 | hCoV-19/Northern Ireland/NIRE-FCE85/2020\|EPI_ISL_4 | ···················· | ···················· | ···················· | ···················· | ···················· | ···················· |
| E | 3459 | hCoV-19/Northern Ireland/NIRE-10360A/2020\|EPI_ISL_ | ···················· | ···················· | ···················· | ···················· | ···················· | ···················· |
| E | 3460 | hCoV-19/Northern Ireland/NIRE-100087/2020\|EPI_ISL_ | ···················· | ···················· | ···················· | ···················· | ···················· | ···················· |
| E | 3461 | hCoV-19/Northern Ireland/NIRE-FD1F4/2020\|EPI_ISL_4 | ···················· | ···················· | ···················· | ···················· | ···················· | ···················· |
| E | 3462 | hCoV-19/Northern Ireland/NIRE-FD2C4/2020\|EPI_ISL_4 | ···················· | ···················· | ···················· | ···················· | ···················· | ···················· |
| E | 3463 | hCoV-19/Northern Ireland/NIRE-103C44/2020\|EPI_ISL_ | ···················· | ···················· | ···················· | ···················· | ···················· | ···················· |
| E | 3464 | hCoV-19/Northern Ireland/NIRE-FD631/2020\|EPI_ISL_4 | ···················· | ·A·················· | ···················· | ···················· | ···················· | ···················· |
| E | 3465 | hCoV-19/Northern Ireland/NIRE-101C19/2020\|EPI_ISL_ | ···················· | ···················· | ···················· | ···················· | ···················· | ···················· |
| E | 3466 | hCoV-19/Northern Ireland/NIRE-FE621/2020\|EPI_ISL_4 | ···················· | ···················· | ···················· | ···················· | ···················· | ···················· |
| E | 3467 | hCoV-19/Northern Ireland/NIRE-103A68/2020\|EPI_ISL_ | ···················· | ···················· | ···················· | ···················· | ···················· | ···················· |
| E | 3468 | hCoV-19/Northern Ireland/NIRE-FE357/2020\|EPI_ISL_4 | ···················· | ···················· | ···················· | ···················· | ···················· | ···················· |
| E | 3469 | hCoV-19/Northern Ireland/NIRE-101AE2/2020\|EPI_ISL_ | ···················· | ···················· | ···················· | ···················· | ···················· | ···················· |
| E | 3470 | hCoV-19/Northern Ireland/NIRE-FE4CD/2020\|EPI_ISL_4 | ···················· | ···················· | ···················· | ···················· | ···················· | ···················· |
| E | 3471 | hCoV-19/Northern Ireland/NIRE-FE2E1/2020\|EPI_ISL_4 | ···················· | ···················· | ···················· | ···················· | ···················· | ···················· |
| E | 3472 | hCoV-19/Northern Ireland/NIRE-101CFB/2020\|EPI_ISL_ | ···················· | ···················· | ···················· | ···················· | ···················· | ···················· |
| E | 3473 | hCoV-19/Northern Ireland/NIRE-FEAF8/2020\|EPI_ISL_4 | ···················· | ···················· | ···················· | ···················· | ···················· | ···················· |
| E | 3474 | hCoV-19/Northern Ireland/NIRE-FEB40/2020\|EPI_ISL_4 | ···················· | ···················· | ···················· | ···················· | ···················· | ···················· |
| E | 3475 | hCoV-19/Northern Ireland/NIRE-FD58F/2020\|EPI_ISL_4 | ···················· | ···················· | ···················· | ···················· | ···················· | ···················· |
| E | 3476 | hCoV-19/Northern Ireland/NIRE-1017EB/2020\|EPI_ISL_ | ···················· | ···················· | ···················· | ···················· | ···················· | ···················· |
| E | 3477 | hCoV-19/Northern Ireland/NIRE-FD710/2020\|EPI_ISL_4 | ···················· | ···················· | ···················· | ···················· | ···················· | ···················· |
| E | 3478 | hCoV-19/Northern Ireland/NIRE-101921/2020\|EPI_ISL_ | ···················· | ···················· | ···················· | ···················· | ···················· | ···················· |
| E | 3479 | hCoV-19/Northern Ireland/NIRE-FE24B/2020\|EPI_ISL_4 | ···················· | ···················· | ···················· | ···················· | ···················· | ···················· |
| E | 3480 | hCoV-19/USA/CA-QRTP-Donor1/2020\|EPI_ISL_490210\|202 | ···················· | ···················· | ···················· | ···················· | ···················· | ···················· |
| E | 3481 | hCoV-19/USA/CA-QRTP-Donor17/2020\|EPI_ISL_490220\|20 | ···················· | ···················· | ···················· | ···················· | ···················· | ···················· |
| E | 3482 | hCoV-19/USA/CA-QRTP-Donor18/2020\|EPI_ISL_490221\|20 | ···················· | ···················· | ···················· | ···················· | ···················· | ···················· |
| E | 3484 | hCoV-19/England/202691845/2020\|EPI_ISL_490226\|2020 | ···················· | ·············A······ | ···················· | ···················· | ···················· | ···················· |
| E | 3485 | hCoV-19/England/202822170/2020\|EPI_ISL_490227\|2020 | ···················· | ·············A······ | ···················· | ···················· | ···················· | ···················· |
| E | 3486 | hCoV-19/England/202822171/2020\|EPI_ISL_490228\|2020 | ···················· | ·············A······ | ···················· | ···················· | ···················· | ···················· |
| E | 3488 | hCoV-19/England/202822194/2020\|EPI_ISL_490250\|2020 | ···················· | ···················· | ···················· | ···················· | ···················· | ···················· |
| E | 3489 | hCoV-19/England/202822195/2020\|EPI_ISL_490251\|2020 | ···················· | ·············A······ | ···················· | ···················· | ···················· | ···················· |
| E | 3490 | hCoV-19/England/202822203/2020\|EPI_ISL_490254\|2020 | ···················· | ·············A······ | ···················· | ···················· | ···················· | ···················· |
| E | 3491 | hCoV-19/Scotland/EDB657/2020\|EPI_ISL_491541\|2020-0 | ···················· | ···················· | ···················· | ···················· | ···················· | ···················· |
| E | 3492 | hCoV-19/Scotland/EDB2547/2020\|EPI_ISL_491550\|2020- | ···················· | ···················· | ···················· | ···················· | ···················· | ···················· |
| E | 3493 | hCoV-19/Scotland/EDB6737/2020\|EPI_ISL_491551\|2020- | ···················· | ···················· | ···················· | ···················· | ···················· | ···················· |
| E | 3494 | hCoV-19/Scotland/EDB690/2020\|EPI_ISL_491552\|2020-0 | ···················· | ···················· | ···················· | ···················· | ···················· | ···················· |
| E | 3495 | hCoV-19/Scotland/EDB6727/2020\|EPI_ISL_491555\|2020- | ···················· | ···················· | ···················· | ···················· | ···················· | ···················· |
| E | 3496 | hCoV-19/Scotland/EDB6443/2020\|EPI_ISL_491556\|2020- | ···················· | ···················· | ···················· | ···················· | ···················· | ···················· |
| E | 3497 | hCoV-19/Scotland/EDB6744/2020\|EPI_ISL_491557\|2020- | ···················· | ···················· | ···················· | ···················· | ···················· | ···················· |

| | | | | | | | | |
|---|---|---|---|---|---|---|---|---|
| E | 3498 hCoV-19/Scotland/EDB614/2020\|EPI_ISL_491558\|2020-0 | ···················· | ···················· | ···················· | ···················· | ···················· | ···················· | |
| E | 3499 hCoV-19/Scotland/EDB1258/2020\|EPI_ISL_491559\|2020- | ···················· | ·A·················· | ···················· | ···················· | ···················· | ···················· | |
| E | 3500 hCoV-19/Scotland/EDB2007/2020\|EPI_ISL_491581\|2020- | ···················· | ···················· | ···················· | ···················· | ···················· | ···················· | |
| E | 3501 hCoV-19/Scotland/EDB1196/2020\|EPI_ISL_491582\|2020- | ···················· | ···················· | ···················· | ···················· | ···················· | ···················· | |
| E | 3502 hCoV-19/Scotland/EDB2143/2020\|EPI_ISL_491583\|2020- | ···················· | ···················· | ···················· | ···················· | ···················· | ···················· | |
| E | 3503 hCoV-19/Scotland/EDB1303/2020\|EPI_ISL_491584\|2020- | ···················· | ···················· | ···················· | ···················· | ···················· | ···················· | |
| E | 3504 hCoV-19/Scotland/EDB1131/2020\|EPI_ISL_491585\|2020- | ···················· | ···················A | ···················· | ···················· | ···················· | ···················· | 1 |
| E | 3505 hCoV-19/Scotland/EDB6734/2020\|EPI_ISL_491587\|2020- | ···················· | ···················· | ···················· | ···················· | ···················· | ···················· | |
| E | 3506 hCoV-19/Scotland/EDB1852/2020\|EPI_ISL_491588\|2020- | ···················· | ···················· | ···················· | ···················· | ···················· | ···················· | |
| E | 3507 hCoV-19/Scotland/EDB1127/2020\|EPI_ISL_491589\|2020- | ···················· | ·A·················· | ···················· | ···················· | ···················· | ···················· | |
| E | 3508 hCoV-19/Scotland/EDB1481/2020\|EPI_ISL_491590\|2020- | ···················· | ·A·················· | ···················· | ···················· | ···················· | ···················· | |
| E | 3509 hCoV-19/Scotland/EDB1922/2020\|EPI_ISL_491591\|2020- | ···················· | ·A·················· | ···················· | ···················· | ···················· | ···················· | |
| E | 3510 hCoV-19/Scotland/EDB1390/2020\|EPI_ISL_491592\|2020- | ···················· | ···················· | ···················· | ···················· | ···················· | ···················· | |
| E | 3511 hCoV-19/Scotland/EDB3582/2020\|EPI_ISL_491593\|2020- | ···················· | ···················· | ···················· | ···················· | ···················· | ···················· | |
| E | 3512 hCoV-19/Scotland/EDB2570/2020\|EPI_ISL_491594\|2020- | ···················· | ···················· | ···················· | ···················· | ···················· | ···················· | |
| E | 3513 hCoV-19/Scotland/EDB1284/2020\|EPI_ISL_491596\|2020- | ···················· | ···················· | ···················· | ···················· | ···················· | ···················· | |
| E | 3514 hCoV-19/Scotland/EDB2550/2020\|EPI_ISL_491597\|2020- | ···················· | ···················· | ···················· | ···················· | ···················· | ···················· | |
| E | 3515 hCoV-19/Scotland/EDB3831/2020\|EPI_ISL_491598\|2020- | ···················· | ···················· | ···················· | ···················· | ···················· | ···················· | |
| E | 3516 hCoV-19/Scotland/EDB2054/2020\|EPI_ISL_491599\|2020- | ···················· | ···················· | ···················· | ···················· | ···················· | ···················· | |
| E | 3517 hCoV-19/England/WSFT-25C138F/2020\|EPI_ISL_492873\|2 | ···················· | ···················· | ···················· | ···················· | ···················· | ···················· | |
| E | 3518 hCoV-19/England/LOND-124C294/2020\|EPI_ISL_492880\|2 | ···················· | ···················· | ···················· | ···················· | ···················· | ···················· | |
| E | 3519 hCoV-19/England/WSFT-25C0F9B/2020\|EPI_ISL_492881\|2 | ···················· | ···················· | ···················· | ···················· | ···················· | ···················· | |
| E | 3520 hCoV-19/England/LOND-124C30A/2020\|EPI_ISL_492882\|2 | ···················· | ···················· | ···················· | ···················· | ···················· | ···················· | |
| E | 3521 hCoV-19/England/WSFT-25C105B/2020\|EPI_ISL_492884\|2 | ···················· | ···················· | ···················· | ···················· | ···················· | ···················· | |
| E | 3522 hCoV-19/England/LOND-124C373/2020\|EPI_ISL_492885\|2 | ···················· | ···················· | ···················· | ···················· | ···················· | ···················· | |
| E | 3523 hCoV-19/England/WSFT-25C13AD/2020\|EPI_ISL_492886\|2 | ···················· | ···················· | ···················· | ···················· | ···················· | ···················· | |
| E | 3524 hCoV-19/England/LOND-124C434/2020\|EPI_ISL_492887\|2 | ···················· | ···················· | ···················· | ···················· | ···················· | ···················· | |
| E | 3525 hCoV-19/England/LOND-124C4CB/2020\|EPI_ISL_492888\|2 | ···················· | ···················· | ···················· | ···················· | ···················· | ···················· | |
| E | 3526 hCoV-19/England/WSFT-25C0DEC/2020\|EPI_ISL_492900\|2 | ···················· | ···················· | ···················· | ···················· | ···················· | ···················· | |
| E | 3527 hCoV-19/England/LOND-124C66B/2020\|EPI_ISL_492902\|2 | ···················· | ···················· | ···················· | ···················· | ···················· | ···················· | |
| E | 3528 hCoV-19/England/WSFT-25C1246/2020\|EPI_ISL_492903\|2 | ···················· | ···················· | ···················· | ···················· | ···················· | ···················· | |
| E | 3529 hCoV-19/England/LOND-124C3A0/2020\|EPI_ISL_492905\|2 | ···················· | ···················· | ···················· | ···················· | ···················· | ···················· | |
| E | 3530 hCoV-19/England/LOND-124C364/2020\|EPI_ISL_492906\|2 | ···················· | ···················· | ···················· | ···················· | ···················· | ···················· | |
| E | 3531 hCoV-19/England/LOND-124C689/2020\|EPI_ISL_492908\|2 | ···················· | ···················· | ···················· | ···················· | ···················· | ···················· | |
| E | 3532 hCoV-19/England/WSFT-25C0DA0/2020\|EPI_ISL_492909\|2 | ···················· | ···················· | ···················· | ···················· | ···················· | ···················· | |
| E | 3533 hCoV-19/England/BRIS-1333EC/2020\|EPI_ISL_495100\|20 | ···················· | ···················· | ···················· | ···················· | ···················· | ···················· | |
| E | 3534 hCoV-19/England/BRIS-185470D/2020\|EPI_ISL_495101\|2 | ···················· | ···················· | ···················· | ···················· | ···················· | ···················· | |
| E | 3535 hCoV-19/England/BRIS-185465B/2020\|EPI_ISL_495102\|2 | ···················· | ···················· | ···················· | ···················· | ···················· | ···················· | |
| E | 3536 hCoV-19/England/BRIS-133461/2020\|EPI_ISL_495103\|20 | ···················· | ···················· | ···················· | ···················· | ···················· | ···················· | |
| E | 3537 hCoV-19/England/BRIS-18547EF/2020\|EPI_ISL_495104\|2 | ···················· | ···················· | ···················· | ···················· | ···················· | ···················· | |
| E | 3538 hCoV-19/England/BRIS-1854767/2020\|EPI_ISL_495107\|2 | ···················· | ···················· | ···················· | ···················· | ···················· | ···················· | |
| E | 3539 hCoV-19/England/BRIS-185462E/2020\|EPI_ISL_495108\|2 | ···················· | ···················· | ···················· | ···················· | ···················· | ···················· | |
| E | 3540 hCoV-19/England/BRIS-13320D/2020\|EPI_ISL_495109\|20 | ···················· | ···················· | ···················· | ···················· | ···················· | ···················· | |
| E | 3541 hCoV-19/England/BRIS-18547A3/2020\|EPI_ISL_495110\|2 | ···················· | ···················· | ···················· | ···················· | ···················· | ···················· | |
| E | 3542 hCoV-19/England/BRIS-1854785/2020\|EPI_ISL_495112\|2 | ···················· | ···················· | ···················· | ···················· | ···················· | ···················· | |
| E | 3543 hCoV-19/England/BRIS-18545F4/2020\|EPI_ISL_495113\|2 | ···················· | ···················· | ···················· | ···················· | ···················· | ···················· | |
| E | 3544 hCoV-19/England/BRIS-1854794/2020\|EPI_ISL_495114\|2 | ···················· | ···················· | ···················· | ···················· | ···················· | ···················· | |
| E | 3545 hCoV-19/England/BRIS-18547FE/2020\|EPI_ISL_495116\|2 | ···················· | ···················· | ···················· | ···················· | ···················· | ···················· | |
| E | 3546 hCoV-19/England/BRIS-1333DD/2020\|EPI_ISL_495117\|20 | ···················· | ···················· | ···················· | ···················· | ···················· | ···················· | |
| E | 3547 hCoV-19/England/BRIS-1331A6/2020\|EPI_ISL_495118\|20 | ···················· | ···················· | ···················· | ···················· | ···················· | ···················· | |
| E | 3548 hCoV-19/England/BRIS-18547B2/2020\|EPI_ISL_495119\|2 | ···················· | ···················· | ···················· | ···················· | ···················· | ···················· | |
| E | 3549 hCoV-19/USA/CA-IGI-0100/2020\|EPI_ISL_495122\|2020-0 | ···················· | ···················· | ···················· | ···················· | ···················· | ···················· | |
| E | 3550 hCoV-19/USA/CA-IGI-0101/2020\|EPI_ISL_495123\|2020-0 | ···················· | ···················· | ···················· | ···················· | ···················· | ···················· | |
| E | 3551 hCoV-19/USA/CA-IGI-0109/2020\|EPI_ISL_495131\|2020-0 | ···················· | ···················· | ···················· | ···················· | ···················· | ···················· | |
| E | 3552 hCoV-19/USA/CA-IGI-0114/2020\|EPI_ISL_495136\|2020-0 | ···················· | ···················· | ···················· | ···················· | ···················· | ···················· | |
| E | 3553 hCoV-19/USA/CA-IGI-0120/2020\|EPI_ISL_495142\|2020-0 | ···················· | ···················· | ···················· | ···················· | ···················· | ···················· | |
| E | 3554 hCoV-19/USA/CA-IGI-0122/2020\|EPI_ISL_495144\|2020-0 | ···················· | ···················· | ···················· | ···················· | ···················· | ···················· | |
| E | 3555 hCoV-19/India/TG-CCMB-L1885/2020\|EPI_ISL_495167\|20 | ···················· | ···················· | ···················· | ···················· | ···················· | ···················· | |
| E | 3557 hCoV-19/India/TG-CCMB-L1893/2020\|EPI_ISL_495169\|20 | ···················· | ···················· | ···················· | ···················· | ···················· | ···················· | |
| E | 3558 hCoV-19/India/TG-CCMB-L1930/2020\|EPI_ISL_495170\|20 | ···················· | ···················· | ···················· | ···················· | ···················· | ···················· | |
| E | 3559 hCoV-19/India/TG-CCMB-L1931/2020\|EPI_ISL_495171\|20 | ···················· | ···················· | ···················· | ···················· | ···················· | ···················· | |
| E | 3560 hCoV-19/India/TG-CCMB-L1934/2020\|EPI_ISL_495172\|20 | ···················· | ···················· | ···················· | ···················· | ···················· | ···················· | |
| E | 3561 hCoV-19/India/TG-CCMB-L1935/2020\|EPI_ISL_495173\|20 | ···················· | ···················· | ···················· | ···················· | ···················· | ···················· | |
| E | 3562 hCoV-19/India/TG-CCMB-L1959/2020\|EPI_ISL_495174\|20 | ···················· | ···················· | ···················· | ···················· | ···················· | ···················· | |
| E | 3563 hCoV-19/India/TG-CCMB-L1963/2020\|EPI_ISL_495175\|20 | ···················· | ···················· | ···················· | ···················· | ···················· | ···················· | |
| E | 3565 hCoV-19/India/TG-CCMB-L1967/2020\|EPI_ISL_495177\|20 | ···················· | ···················· | ···················· | ···················· | ···················· | ···················· | |
| E | 3566 hCoV-19/India/TG-CCMB-L1973/2020\|EPI_ISL_495178\|20 | ···················· | ···················· | ···················· | ···················· | ···················· | ···················· | |
| E | 3567 hCoV-19/India/TG-CCMB-L1975/2020\|EPI_ISL_495179\|20 | ···················· | ···················· | ···················· | ···················· | ···················· | ···················· | |
| E | 3568 hCoV-19/India/TG-CCMB-L1999/2020\|EPI_ISL_495180\|20 | ···················· | ···················· | ···················· | ···················· | ···················· | ···················· | |
| E | 3569 hCoV-19/India/TG-CCMB-M106/2020\|EPI_ISL_495181\|202 | ···················· | ···················· | ···················· | ···················· | ···················· | ···················· | |
| E | 3570 hCoV-19/India/TG-CCMB-M116/2020\|EPI_ISL_495182\|202 | ···················· | ···················· | ···················· | ···················· | ···················· | ···················· | |
| E | 3571 hCoV-19/India/TG-CCMB-M118/2020\|EPI_ISL_495183\|202 | ···················· | ···················· | ···················· | ···················· | ···················· | ···················· | |
| E | 3572 hCoV-19/India/TG-CCMB-M135/2020\|EPI_ISL_495184\|202 | ···················· | ···················· | ···················· | ···················· | ···················· | ···················· | |
| E | 3573 hCoV-19/India/TG-CCMB-M14/2020\|EPI_ISL_495185\|2020 | ···················· | ···················· | ···················· | ···················· | ···················· | ···················· | |
| E | 3574 hCoV-19/India/TG-CCMB-M151/2020\|EPI_ISL_495186\|202 | ···················· | ···················· | ···················· | ···················· | ···················· | ···················· | |
| E | 3575 hCoV-19/India/TG-CCMB-M152/2020\|EPI_ISL_495187\|202 | ···················· | ···················· | ···················· | ···················· | ···················· | ···················· | |
| E | 3576 hCoV-19/India/TG-CCMB-M170/2020\|EPI_ISL_495188\|202 | ···················· | ···················· | ···················· | ···················· | ···················· | ···················· | |
| E | 3577 hCoV-19/Germany/BY-UHR-3ZK/2020\|EPI_ISL_513300\|202 | ···················· | ···················· | ···················· | ···················· | ···················· | ···················· | |
| E | 3578 hCoV-19/Germany/BY-UHR-4ZK/2020\|EPI_ISL_513301\|202 | ···················· | ···················· | ···················· | ···················· | ···················· | ···················· | |
| E | 3579 hCoV-19/Germany/BY-UHR-5ZK/2020\|EPI_ISL_513302\|202 | ···················· | ···················· | ···················· | ···················· | ···················· | ···················· | |

| | | | | | | | |
|---|---|---|---|---|---|---|---|
| E | 3580 hCoV-19/Germany/BY-UHR-6ZK/2020\|EPI_ISL_513303\|202 | ···················· | ···················· | ····················· | ···················· | ···················· | ···················· |
| E | 3581 hCoV-19/Germany/BY-UHR-7ZK/2020\|EPI_ISL_513304\|202 | ···················· | ···················· | ····················· | ···················· | ···················· | ···················· |
| E | 3582 hCoV-19/Germany/BY-UHR-8ZK/2020\|EPI_ISL_513305\|202 | ···················· | ···················· | ····················· | ···················· | ···················· | ···················· |
| E | 3583 hCoV-19/Germany/BY-UHR-9ZK/2020\|EPI_ISL_513306\|202 | ···················· | ···················· | ····················· | ···················· | ···················· | ···················· |
| E | 3584 hCoV-19/Germany/BY-UHR-10ZK/2020\|EPI_ISL_513307\|20 | ···················· | ···················· | ····················· | ···················· | ···················· | ···················· |
| E | 3585 hCoV-19/Canada/NB-202/2020\|EPI_ISL_479625\|2020-03- | ···················· | ···················· | ····················· | ···················· | ···················· | ···················· |
| E | 3586 hCoV-19/Canada/NB-204/2020\|EPI_ISL_479626\|2020-03- | ···················· | ···················· | ····················· | ···················· | ···················· | ···················· |
| E | 3587 hCoV-19/Canada/NB-206/2020\|EPI_ISL_479627\|2020-03- | ···················· | ···················· | ····················· | ···················· | ···················· | ···················· |
| E | 3588 hCoV-19/Canada/NB-209/2020\|EPI_ISL_479628\|2020-03- | ···················· | ···················· | ····················· | ···················· | ···················· | ···················· |
| E | 3589 hCoV-19/Canada/NB-213/2020\|EPI_ISL_479629\|2020-03- | ···················· | ···················· | ····················· | ···················· | ···················· | ···················· |
| E | 3590 hCoV-19/Canada/NB-214/2020\|EPI_ISL_479630\|2020-03- | ···················· | ···················· | ····················· | ···················· | ···················· | ···················· |
| E | 3591 hCoV-19/Canada/NB-215/2020\|EPI_ISL_479631\|2020-03- | ···················· | ···················· | ····················· | ···················· | ···················· | ···················· |
| E | 3592 hCoV-19/Canada/NB-216/2020\|EPI_ISL_479632\|2020-03- | ···················· | ···················· | ····················· | ···················· | ···················· | ···················· |
| E | 3593 hCoV-19/Canada/NB-217/2020\|EPI_ISL_479633\|2020-03- | ···················· | ···················· | ····················· | ···················· | ···················· | ···················· |
| E | 3594 hCoV-19/Canada/NB-218/2020\|EPI_ISL_479634\|2020-03- | ···················· | ···················· | ····················· | ···················· | ···················· | ···················· |
| E | 3595 hCoV-19/Canada/NB-219/2020\|EPI_ISL_479635\|2020-03- | ···················· | ···················· | ····················· | ···················· | ···················· | ···················· |
| E | 3596 hCoV-19/Canada/NB-220/2020\|EPI_ISL_479636\|2020-03- | ···················· | ···················· | ····················· | ···················· | ···················· | ···················· |
| E | 3597 hCoV-19/Canada/NB-221/2020\|EPI_ISL_479637\|2020-03- | ···················· | ···················· | ····················· | ···················· | ···················· | ···················· |
| E | 3598 hCoV-19/Canada/NB-222/2020\|EPI_ISL_479638\|2020-03- | ···················· | ···················· | ····················· | ···················· | ···················· | ···················· |
| E | 3599 hCoV-19/Canada/NB-223/2020\|EPI_ISL_479639\|2020-03- | ···················· | ·············A······ | ····················· | ···················· | ···················· | ···················· |
| E | 3600 hCoV-19/Canada/NB-224/2020\|EPI_ISL_479640\|2020-03- | ···················· | ···················· | ····················· | ···················· | ···················· | ···················· |
| E | 3601 hCoV-19/Canada/NB-225/2020\|EPI_ISL_479641\|2020-03- | ···················· | ···················· | ····················· | ···················· | ···················· | ···················· |
| E | 3602 hCoV-19/Canada/NB-226/2020\|EPI_ISL_479642\|2020-03- | ···················· | ···················· | ····················· | ···················· | ···················· | ···················· |
| E | 3603 hCoV-19/Canada/NB-228/2020\|EPI_ISL_479643\|2020-03- | ···················· | ···················· | ····················· | ···················· | ···················· | ···················· |
| E | 3604 hCoV-19/Canada/NB-231/2020\|EPI_ISL_479644\|2020-03- | ···················· | ···················· | ····················· | ···················· | ···················· | ···················· |
| E | 3605 hCoV-19/Canada/NB-232/2020\|EPI_ISL_479645\|2020-03- | ···················· | ···················· | ····················· | ···················· | ···················· | ···················· |
| E | 3606 hCoV-19/Canada/NB-233/2020\|EPI_ISL_479646\|2020-03- | ···················· | ···················· | ····················· | ···················· | ···················· | ···················· |
| E | 3607 hCoV-19/Canada/NB-234/2020\|EPI_ISL_479647\|2020-03- | ···················· | ···················· | ····················· | ···················· | ···················· | ···················· |
| E | 3608 hCoV-19/Canada/NB-235/2020\|EPI_ISL_479648\|2020-03- | ···················· | ···················· | ····················· | ···················· | ···················· | ···················· |
| E | 3609 hCoV-19/Canada/NB-236/2020\|EPI_ISL_479649\|2020-03- | ···················· | ···················· | ····················· | ···················· | ···················· | ···················· |
| E | 3610 hCoV-19/Canada/NB-237/2020\|EPI_ISL_479650\|2020-03- | ···················· | ···················· | ····················· | ···················· | ···················· | ···················· |
| E | 3611 hCoV-19/Canada/NB-238/2020\|EPI_ISL_479651\|2020-03- | ···················· | ···················· | ····················· | ···················· | ···················· | ···················· |
| E | 3615 hCoV-19/Australia/VIC2076/2020\|EPI_ISL_480616\|2020 | ···················· | ···················· | ····················· | ···················· | ···················· | ···················· |
| E | 3623 hCoV-19/Australia/VIC1891/2020\|EPI_ISL_480641\|2020 | ···················· | ···················· | ····················· | ···················· | ···················· | ···················· |
| E | 3627 hCoV-19/England/BRIS-12D813/2020\|EPI_ISL_481900\|20 | ···················· | ···················· | ····················· | ···················· | ···················· | ···················· |
| E | 3628 hCoV-19/England/BRIS-12D95C/2020\|EPI_ISL_481901\|20 | ···················· | ···················· | ····················· | ···················· | ···················· | ···················· |
| E | 3629 hCoV-19/England/BRIS-12CB75/2020\|EPI_ISL_481902\|20 | ···················· | ···················· | ····················· | ···················· | ···················· | ···················· |
| E | 3630 hCoV-19/England/BRIS-12DA4A/2020\|EPI_ISL_481903\|20 | ···················· | ···················· | ····················· | ···················· | ···················· | ···················· |
| E | 3631 hCoV-19/England/BRIS-12D075/2020\|EPI_ISL_481904\|20 | ···················· | ···················· | ····················· | ···················· | ···················· | ···················· |
| E | 3632 hCoV-19/England/BRIS-12D761/2020\|EPI_ISL_481905\|20 | ···················· | ···················· | ····················· | ···················· | ···················· | ···················· |
| E | 3633 hCoV-19/England/BRIS-12CB39/2020\|EPI_ISL_481906\|20 | ···················· | ···················· | ····················· | ···················· | ···················· | ···················· |
| E | 3634 hCoV-19/England/BRIS-12D206/2020\|EPI_ISL_481907\|20 | ···················· | ···················· | ····················· | ···················· | ···················· | ···················· |
| E | 3635 hCoV-19/England/BRIS-12CC36/2020\|EPI_ISL_481909\|20 | ···················· | ···················· | ····················· | ···················· | ···················· | ···················· |
| E | 3636 hCoV-19/England/BRIS-12CFF1/2020\|EPI_ISL_481910\|20 | ···················· | ···················· | ····················· | ···················· | ···················· | ···················· |
| E | 3637 hCoV-19/England/BRIS-12CE8B/2020\|EPI_ISL_481911\|20 | ···················· | ···················· | ····················· | ···················· | ···················· | ···················· |
| E | 3638 hCoV-19/England/BRIS-12D8B9/2020\|EPI_ISL_481912\|20 | ···················· | ···················· | ····················· | ···················· | ···················· | ···················· |
| E | 3639 hCoV-19/England/BRIS-12D558/2020\|EPI_ISL_481913\|20 | ···················· | ···················· | ····················· | ···················· | ···················· | ···················· |
| E | 3640 hCoV-19/England/BRIS-12CF2E/2020\|EPI_ISL_481914\|20 | ···················· | ···················· | ····················· | ···················· | ···················· | ···················· |
| E | 3641 hCoV-19/England/BRIS-12D1EB/2020\|EPI_ISL_481915\|20 | ···················· | ···················· | ····················· | ···················· | ···················· | ···················· |
| E | 3642 hCoV-19/England/BRIS-12D8E6/2020\|EPI_ISL_481917\|20 | ···················· | ···················· | ····················· | ···················· | ···················· | ···················· |
| E | 3643 hCoV-19/England/BRIS-12D0EE/2020\|EPI_ISL_481918\|20 | ···················· | ···················· | ····················· | ···················· | ···················· | ···················· |
| E | 3644 hCoV-19/England/BRIS-12D093/2020\|EPI_ISL_481919\|20 | ···················· | ···················· | ····················· | ···················· | ···················· | ···················· |
| E | 3645 hCoV-19/England/BRIS-12D109/2020\|EPI_ISL_481920\|20 | ···················· | ····A··············· | ····················· | ···················· | ···················· | ···················· |
| E | 3646 hCoV-19/England/BRIS-12D066/2020\|EPI_ISL_481921\|20 | ···················· | ···················· | ····················· | ···················· | ···················· | ···················· |
| E | 3647 hCoV-19/England/BRIS-12CFC4/2020\|EPI_ISL_481922\|20 | ···················· | ···················· | ····················· | ···················· | ···················· | ···················· |
| E | 3648 hCoV-19/England/BRIS-12CC27/2020\|EPI_ISL_481924\|20 | ···················· | ···················· | ····················· | ···················· | ···················· | ···················· |
| E | 3649 hCoV-19/England/BRIS-12D312/2020\|EPI_ISL_481925\|20 | ···················· | ···················· | ····················· | ···················· | ···················· | ···················· |
| E | 3650 hCoV-19/England/BRIS-12D51C/2020\|EPI_ISL_481926\|20 | ···················· | ···················· | ····················· | ···················· | ···················· | ···················· |
| E | 3651 hCoV-19/England/BRIS-12D400/2020\|EPI_ISL_481927\|20 | ···················· | ···················· | ····················· | ···················· | ···················· | ···················· |
| E | 3652 hCoV-19/England/BRIS-12D01B/2020\|EPI_ISL_481929\|20 | ···················· | ···················· | ···T················· | ···················· | ···················· | ···················· |
| E | 3653 hCoV-19/England/BRIS-12D682/2020\|EPI_ISL_481930\|20 | ···················· | ···················· | ····················· | ···················· | ···················· | ···················· |
| E | 3654 hCoV-19/England/BRIS-12DAE0/2020\|EPI_ISL_481931\|20 | ···················· | ···················· | ····················· | ···················· | ···················· | ···················· |
| E | 3655 hCoV-19/England/BRIS-12CBC0/2020\|EPI_ISL_481932\|20 | ···················· | ···················· | ····················· | ···················· | ···················· | ···················· |
| E | 3656 hCoV-19/England/BRIS-12CE9A/2020\|EPI_ISL_481933\|20 | ···················· | ···················· | ····················· | ···················· | ···················· | ···················· |
| E | 3657 hCoV-19/England/BRIS-12D549/2020\|EPI_ISL_481934\|20 | ···················· | ···················· | ····················· | ···················· | ···················· | ···················· |
| E | 3658 hCoV-19/England/BRIS-12CA78/2020\|EPI_ISL_481935\|20 | ···················· | ···················· | ····················· | ···················· | ···················· | ···················· |
| E | 3659 hCoV-19/England/BRIS-12CD7F/2020\|EPI_ISL_481936\|20 | ···················· | ···················· | ····················· | ···················· | ···················· | ···················· |
| E | 3660 hCoV-19/England/BRIS-12CDAC/2020\|EPI_ISL_481938\|20 | ···················· | ···················· | ····················· | ···················· | ···················· | ···················· |
| E | 3661 hCoV-19/England/BRIS-12CEA9/2020\|EPI_ISL_481939\|20 | ···················· | ···················· | ····················· | ···················· | ···················· | ···················· |
| E | 3662 hCoV-19/England/BRIS-12DA1D/2020\|EPI_ISL_481940\|20 | ···················· | ···················· | ····················· | ···················· | ···················· | ···················· |
| E | 3663 hCoV-19/England/BRIS-12D3C7/2020\|EPI_ISL_481941\|20 | ···················· | ···················· | ····················· | ···················· | ···················· | ···················· |
| E | 3664 hCoV-19/England/BRIS-12CDCA/2020\|EPI_ISL_481942\|20 | ···················· | ···················· | ····················· | ···················· | ···················· | ···················· |
| E | 3665 hCoV-19/England/BRIS-12D00C/2020\|EPI_ISL_481943\|20 | ···················· | ···················· | ····················· | ···················· | ···················· | ···················· |
| E | 3666 hCoV-19/England/BRIS-12CFD3/2020\|EPI_ISL_481945\|20 | ···················· | ···················· | ····················· | ···················· | ···················· | ···················· |
| E | 3667 hCoV-19/England/BRIS-12D628/2020\|EPI_ISL_481946\|20 | ···················· | ···················· | ····················· | ···················· | ···················· | ···················· |
| E | 3668 hCoV-19/England/BRIS-12D0B1/2020\|EPI_ISL_481947\|20 | ···················· | ···················· | ····················· | ···················· | ···················· | ···················· |
| E | 3669 hCoV-19/England/BRIS-12CFA6/2020\|EPI_ISL_481948\|20 | ···················· | ···················· | ····················· | ···················· | ···················· | ···················· |
| E | 3670 hCoV-19/England/BRIS-12CBB1/2020\|EPI_ISL_481949\|20 | ···················· | ···················· | ····················· | ···················· | ···················· | ···················· |
| E | 3671 hCoV-19/England/BRIS-12D39A/2020\|EPI_ISL_481950\|20 | ···················· | ···················· | ····················· | ···················· | ···················· | ···················· |
| E | 3672 hCoV-19/England/BRIS-12D87D/2020\|EPI_ISL_481951\|20 | ···················· | ···················· | ····················· | A··················· | ···················· | ···················· |

| | | | | | | | | |
|---|---|---|---|---|---|---|---|---|
| E | 3673 | hCoV-19/England/BRIS-12D45B/2020\|EPI_ISL_481952\|20 | ···················· | ···················· | ····················· | ···················· | ···················· | ···················· |
| E | 3674 | hCoV-19/England/BRIS-12CAA5/2020\|EPI_ISL_481954\|20 | ···················· | ···················· | ····················· | ···················· | ···················· | ···················· |
| E | 3675 | hCoV-19/England/BRIS-12CC18/2020\|EPI_ISL_481957\|20 | ···················· | ···················· | ····················· | ···················· | ···················· | ···················· |
| E | 3676 | hCoV-19/England/BRIS-12CCBE/2020\|EPI_ISL_481958\|20 | ···················· | ···················· | ····················· | ···················· | ···················· | ···················· |
| E | 3677 | hCoV-19/England/BRIS-12CA3C/2020\|EPI_ISL_481960\|20 | ···················· | ···················· | ····················· | ···················· | ···················· | ···················· |
| E | 3678 | hCoV-19/England/BRIS-12D637/2020\|EPI_ISL_481961\|20 | ···················· | ···················· | ····················· | ···················· | ···················· | ···················· |
| E | 3679 | hCoV-19/England/BRIS-12CFE2/2020\|EPI_ISL_481962\|20 | ···················· | ···················· | ····················· | ···················· | ···················· | ···················· |
| E | 3680 | hCoV-19/England/BRIS-12D0C0/2020\|EPI_ISL_481964\|20 | ···················· | ···················· | ····················· | ···················· | ···················· | ···················· |
| E | 3681 | hCoV-19/England/BRIS-12CCAF/2020\|EPI_ISL_481966\|20 | ···················· | ···················· | ····················· | ···················· | ···················· | ···················· |
| E | 3682 | hCoV-19/England/BRIS-12DDF6/2020\|EPI_ISL_481968\|20 | ···················· | ···················· | ····················· | ···················· | ···················· | ···················· |
| E | 3683 | hCoV-19/England/BRIS-12E3B7/2020\|EPI_ISL_481969\|20 | ···················· | ···················· | ····················· | ···················· | ···················· | ···················· |
| E | 3684 | hCoV-19/England/BRIS-12E311/2020\|EPI_ISL_481971\|20 | ···················· | ···················· | ····················· | ···················· | ···················· | ···················· |
| E | 3685 | hCoV-19/England/BRIS-12E320/2020\|EPI_ISL_481972\|20 | ···················· | ···················· | ····················· | ···················· | ···················· | ···················· |
| E | 3686 | hCoV-19/England/BRIS-12E144/2020\|EPI_ISL_481973\|20 | ···················· | ···················· | ····················· | ···················· | ···················· | ···················· |
| E | 3687 | hCoV-19/England/BRIS-12E33F/2020\|EPI_ISL_481974\|20 | ···················· | ···················· | ····················· | ···················· | ···················· | ···················· |
| E | 3688 | hCoV-19/England/BRIS-12DD9C/2020\|EPI_ISL_481975\|20 | ···················· | ···················· | ····················· | ···················· | ···················· | ···················· |
| E | 3689 | hCoV-19/England/BRIS-12E223/2020\|EPI_ISL_481976\|20 | ···················· | ···················· | ····················· | ···················· | ···················· | ···················· |
| E | 3690 | hCoV-19/England/BRIS-12DF87/2020\|EPI_ISL_481977\|20 | ···················· | ···················· | ····················· | ···················· | ···················· | ···················· |
| E | 3691 | hCoV-19/England/GSTT-265DFB0/2020\|EPI_ISL_484134\|2 | ···················· | ···················· | ····················· | ···················· | ···················· | ···················· |
| E | 3692 | hCoV-19/England/GSTT-265DFDE/2020\|EPI_ISL_484136\|2 | ···················· | ···················· | ····················· | ···················· | ···················· | ···················· |
| E | 3693 | hCoV-19/England/GSTT-265E070/2020\|EPI_ISL_484145\|2 | ···················· | ···················· | ····················· | ···················· | ···················· | ···················· |
| E | 3694 | hCoV-19/England/GSTT-265E0CB/2020\|EPI_ISL_484150\|2 | ···················· | ···················· | ····················· | ···················· | ···················· | ···················· |
| E | 3695 | hCoV-19/England/GSTT-265E0E9/2020\|EPI_ISL_484152\|2 | ···················· | ···················· | ····················· | ···················· | ···················· | ···················· |
| E | 3696 | hCoV-19/England/GSTT-265E201/2020\|EPI_ISL_484170\|2 | ···················· | ···················· | ····················· | ···················· | ···················· | ···················· |
| E | 3697 | hCoV-19/England/GSTT-265E24D/2020\|EPI_ISL_484174\|2 | ···················· | ···················· | ····················· | ···················· | ···················· | ···················· |
| E | 3698 | hCoV-19/England/GSTT-265E34A/2020\|EPI_ISL_484189\|2 | ···················· | ···················· | ····················· | ···················· | ···················· | ···················· |
| E | 3699 | hCoV-19/England/GSTT-265E368/2020\|EPI_ISL_484191\|2 | ···················· | ···················· | ····················· | ···················· | ···················· | ···················· |
| E | 3700 | hCoV-19/Northern Ireland/NIRE-FE339/2020\|EPI_ISL_4 | ···················· | ···················· | ····················· | ···················· | ···················· | ···················· |
| E | 3701 | hCoV-19/Northern Ireland/NIRE-FD525/2020\|EPI_ISL_4 | ···················· | ···················· | ····················· | ···················· | ···················· | ···················· |
| E | 3702 | hCoV-19/Northern Ireland/NIRE-101815/2020\|EPI_ISL_ | ···················· | ···················· | ····················· | ···················· | ···················· | ···················· |
| E | 3703 | hCoV-19/Northern Ireland/NIRE-103BED/2020\|EPI_ISL_ | ···················· | ···················· | ····················· | ···················· | ···················· | ···················· |
| E | 3704 | hCoV-19/Northern Ireland/NIRE-102E21/2020\|EPI_ISL_ | ···················· | ···················· | ····················· | ···················· | ···················· | ···················· |
| E | 3705 | hCoV-19/Northern Ireland/NIRE-103840/2020\|EPI_ISL_ | ···················· | ···················· | ····················· | ···················· | ···················· | ···················· |
| E | 3706 | hCoV-19/Northern Ireland/NIRE-FD455/2020\|EPI_ISL_4 | ···················· | ···················· | ····················· | ···················· | ···················· | ···················· |
| E | 3707 | hCoV-19/Northern Ireland/NIRE-FEB7D/2020\|EPI_ISL_4 | ···················· | ···················· | ····················· | ···················· | ···················· | ···················· |
| E | 3708 | hCoV-19/Northern Ireland/NIRE-10188E/2020\|EPI_ISL_ | ···················· | ···················· | ····················· | ···················· | ···················· | ···················· |
| E | 3709 | hCoV-19/Northern Ireland/NIRE-10001E/2020\|EPI_ISL_ | ···················· | ···················· | ····················· | ···················· | ···················· | ···················· |
| E | 3710 | hCoV-19/Northern Ireland/NIRE-FE445/2020\|EPI_ISL_4 | ···················· | ···················· | ····················· | ···················· | ···················· | ···················· |
| E | 3711 | hCoV-19/Northern Ireland/NIRE-100096/2020\|EPI_ISL_ | ···················· | ···················· | ····················· | ···················· | ···················· | ···················· |
| E | 3712 | hCoV-19/Northern Ireland/NIRE-1043A8/2020\|EPI_ISL_ | ···················· | ···················· | ····················· | ···················· | ···················· | ···················· |
| E | 3713 | hCoV-19/USA/CA-QRTP-Donor2/2020\|EPI_ISL_490211\|202 | ···················· | ···················· | ····················· | ···················· | ···················· | ···················· |
| E | 3714 | hCoV-19/USA/CA-QRTP-Donor3/2020\|EPI_ISL_490212\|202 | ···················· | ···················· | ····················· | ···················· | ···················· | ···················· |
| E | 3715 | hCoV-19/USA/CA-QRTP-Donor4/2020\|EPI_ISL_490213\|202 | ···················· | ···················· | ····················· | ···················· | ···················· | ···················· |
| E | 3716 | hCoV-19/USA/CA-QRTP-Donor5/2020\|EPI_ISL_490214\|202 | ···················· | ···················· | ····················· | ···················· | ···················· | ···················· |
| E | 3717 | hCoV-19/USA/CA-QRTP-Donor6/2020\|EPI_ISL_490215\|202 | ···················· | ···················· | ····················· | ···················· | ···················· | ···················· |
| E | 3718 | hCoV-19/USA/CA-QRTP-Donor10/2020\|EPI_ISL_490216\|20 | ···················· | ···················· | ····················· | ···················· | ···················· | ···················· |
| E | 3719 | hCoV-19/USA/CA-QRTP-Donor13/2020\|EPI_ISL_490217\|20 | ···················· | ···················· | ····················· | ···················· | ···················· | ···················· |
| E | 3720 | hCoV-19/USA/CA-QRTP-Donor15/2020\|EPI_ISL_490219\|20 | ···················· | ···················· | ····················· | ···················· | ···················· | ···················· |
| E | 3721 | hCoV-19/England/202822175/2020\|EPI_ISL_490232\|2020 | ···················· | ·············A······ | ····················· | ···················· | ···················· | ···················· |
| E | 3722 | hCoV-19/England/202822176/2020\|EPI_ISL_490233\|2020 | ···················· | ·············A······ | ····················· | ···················· | ···················· | ···················· |
| E | 3723 | hCoV-19/England/202822177/2020\|EPI_ISL_490234\|2020 | ···················· | ···················· | ····················· | ···················· | ···················· | ···················· |
| E | 3724 | hCoV-19/England/202822178/2020\|EPI_ISL_490235\|2020 | ···················· | ·············A······ | ····················· | ···················· | ···················· | ···················· |
| E | 3725 | hCoV-19/England/202822180/2020\|EPI_ISL_490236\|2020 | ···················· | ·············A······ | ····················· | ···················· | ···················· | ···················· |
| E | 3726 | hCoV-19/England/202822181/2020\|EPI_ISL_490237\|2020 | ···················· | ·············A······ | ····················· | ···················· | ···················· | ···················· |
| E | 3727 | hCoV-19/England/202822182/2020\|EPI_ISL_490238\|2020 | ···················· | ·············A······ | ····················· | ···················· | ···················· | ···················· |
| E | 3728 | hCoV-19/England/202822183/2020\|EPI_ISL_490239\|2020 | ···················· | ·············A······ | ····················· | ···················· | ···················· | ···················· |
| E | 3729 | hCoV-19/England/202822184/2020\|EPI_ISL_490240\|2020 | ···················· | ·············A······ | ····················· | ···················· | ···················· | ···················· |
| E | 3730 | hCoV-19/England/202822185/2020\|EPI_ISL_490241\|2020 | ···················· | ·············A······ | ····················· | ···················· | ···················· | ···················· |
| E | 3731 | hCoV-19/England/202822186/2020\|EPI_ISL_490242\|2020 | ···················· | ·············A······ | ····················· | ···················· | ···················· | ···················· |
| E | 3732 | hCoV-19/England/202822187/2020\|EPI_ISL_490243\|2020 | ···················· | ·············A······ | ····················· | ···················· | ···················· | ···················· |
| E | 3733 | hCoV-19/England/202822188/2020\|EPI_ISL_490244\|2020 | ···················· | ·············A······ | ····················· | ···················· | ···················· | ···················· |
| E | 3734 | hCoV-19/England/202822189/2020\|EPI_ISL_490245\|2020 | ···················· | ·············A······ | ····················· | ···················· | ···················· | ···················· |
| E | 3735 | hCoV-19/England/202822190/2020\|EPI_ISL_490246\|2020 | ···················· | ·············A······ | ····················· | ···················· | ···················· | ···················· |
| E | 3736 | hCoV-19/England/202822191/2020\|EPI_ISL_490247\|2020 | ···················· | ···················· | ····················· | ···················· | ···················· | ···················· |
| E | 3737 | hCoV-19/England/202822193/2020\|EPI_ISL_490249\|2020 | ···················· | ···················· | ····················· | ···················· | ···················· | ···················· |
| E | 3738 | hCoV-19/Scotland/EDB2254/2020\|EPI_ISL_491542\|2020- | ···················· | ···················· | ····················· | ···················· | ···················· | ···················· |
| E | 3739 | hCoV-19/Scotland/EDB1204/2020\|EPI_ISL_491544\|2020- | ···················· | ···················· | ····················· | ···················· | ···················· | ···················· |
| E | 3740 | hCoV-19/Scotland/EDB969/2020\|EPI_ISL_491546\|2020-0 | ···················· | ···················· | ····················· | ···················· | ···················· | ···················· |
| E | 3741 | hCoV-19/Scotland/EDB6739/2020\|EPI_ISL_491549\|2020- | ···················· | ···················· | ····················· | ···················· | ···················· | ···················· |
| E | 3742 | hCoV-19/Scotland/EDB6730/2020\|EPI_ISL_491560\|2020- | ···················· | ·A·················· | ····················· | ···················· | ···················· | ···················· |
| E | 3743 | hCoV-19/Scotland/EDB1332/2020\|EPI_ISL_491561\|2020- | ···················· | ·A·················· | ····················· | ···················· | ···················· | ···················· |
| E | 3744 | hCoV-19/Scotland/EDB6733/2020\|EPI_ISL_491562\|2020- | ···················· | ···················· | ····················· | ···················· | ···················· | ···················· |
| E | 3745 | hCoV-19/Scotland/EDB6736/2020\|EPI_ISL_491563\|2020- | ···················· | ·A·················· | ····················· | ···················· | ···················· | ···················· |
| E | 3746 | hCoV-19/Scotland/EDB1243/2020\|EPI_ISL_491564\|2020- | ···················· | ···················· | ····················· | ···················· | ···················· | ···················· |
| E | 3747 | hCoV-19/Scotland/EDB2008/2020\|EPI_ISL_491565\|2020- | ···················· | ···················· | ····················· | ···················· | ···················· | ···················· |
| E | 3748 | hCoV-19/Scotland/EDB1926/2020\|EPI_ISL_491566\|2020- | ···················· | ···················· | ····················· | ···················· | ···················· | ···················· |
| E | 3749 | hCoV-19/Scotland/EDB2350/2020\|EPI_ISL_491567\|2020- | ···················· | ···················· | ····················· | ···················· | ···················· | ···················· |
| E | 3750 | hCoV-19/Scotland/EDB1035/2020\|EPI_ISL_491569\|2020- | ···················· | ···················· | ····················· | ···················· | ···················· | ···················· |
| E | 3751 | hCoV-19/Scotland/EDB2353/2020\|EPI_ISL_491570\|2020- | ···················· | ·A·················· | ····················· | ···················· | ···················· | ···················· |
| E | 3752 | hCoV-19/Scotland/EDB2566/2020\|EPI_ISL_491571\|2020- | ···················· | ···················· | ····················· | ···················· | ···················· | ···················· |

| | | | | | | | | | |
|---|---|---|---|---|---|---|---|---|---|
| E | 3753 | hCoV-19/Scotland/EDB3876/2020\|EPI_ISL_491573\|2020- | ···················· | ···················· | ···················· | ···················· | ···················· | ···················· | |
| E | 3754 | hCoV-19/Scotland/EDB6724/2020\|EPI_ISL_491574\|2020- | ···················· | ···················· | ···················· | ···················· | ···················· | ···················· | |
| E | 3755 | hCoV-19/Scotland/EDB2116/2020\|EPI_ISL_491575\|2020- | ···················· | ···················A | ···················· | ···················· | ···················· | ···················· | 1 |
| E | 3756 | hCoV-19/Scotland/EDB1002/2020\|EPI_ISL_491576\|2020- | ···················· | ···················· | ···················· | ···················· | ···················· | ···················· | |
| E | 3757 | hCoV-19/Scotland/EDB6447/2020\|EPI_ISL_491577\|2020- | ···················· | ···················· | ···················· | ···················· | ···················· | ···················· | |
| E | 3758 | hCoV-19/Scotland/EDB656/2020\|EPI_ISL_491578\|2020-0 | ···················· | ···················· | ···················· | ···················· | ···················· | ···················· | |
| E | 3759 | hCoV-19/Scotland/EDB2133/2020\|EPI_ISL_491580\|2020- | ···················· | ···················· | ···················· | ···················· | ···················· | ···················· | |
| E | 3760 | hCoV-19/England/BRIS-12FD8B/2020\|EPI_ISL_492600\|20 | ···················· | ···················· | ···················· | ···················· | ···················· | ···················· | |
| E | 3761 | hCoV-19/England/BRIS-130992/2020\|EPI_ISL_492601\|20 | ···················· | ···················· | ···················· | ···················· | ···················· | ···················· | |
| E | 3762 | hCoV-19/England/BRIS-1301A9/2020\|EPI_ISL_492602\|20 | ···················· | ···················· | ···················· | ···················· | ···················· | ···················· | |
| E | 3763 | hCoV-19/England/BRIS-130A80/2020\|EPI_ISL_492604\|20 | ···················· | ···················· | ···················· | ···················· | ···················· | ···················· | |
| E | 3764 | hCoV-19/England/BRIS-130394/2020\|EPI_ISL_492605\|20 | ···················· | ···················· | ···················· | ···················· | ···················· | ···················· | |
| E | 3765 | hCoV-19/England/BRIS-130BE7/2020\|EPI_ISL_492606\|20 | ···················· | ···················· | ···················· | ···················· | ···················· | ···················· | |
| E | 3766 | hCoV-19/England/BRIS-130CA8/2020\|EPI_ISL_492607\|20 | ···················· | ···················· | ···················· | ···················· | ···················· | ···················· | |
| E | 3767 | hCoV-19/England/BRIS-1303D0/2020\|EPI_ISL_492608\|20 | ···················· | ···················· | ···················· | ···················· | ···················· | ···················· | |
| E | 3768 | hCoV-19/England/BRIS-130491/2020\|EPI_ISL_492609\|20 | ···················· | ···················· | ···················· | ···················· | ···················· | ···················· | |
| E | 3769 | hCoV-19/England/BRIS-1309B0/2020\|EPI_ISL_492610\|20 | ···················· | ···················· | ···················· | ···················· | ···················· | ···················· | |
| E | 3770 | hCoV-19/England/BRIS-130604/2020\|EPI_ISL_492611\|20 | ···················· | ···················· | ···················· | ···················· | ···················· | ···················· | |
| E | 3771 | hCoV-19/England/BRIS-1306F5/2020\|EPI_ISL_492612\|20 | ···················· | ···················· | ···················· | ···················· | ···················· | ···················· | |
| E | 3772 | hCoV-19/England/BRIS-130710/2020\|EPI_ISL_492613\|20 | ···················· | ···················· | ···················· | ···················· | ···················· | ···················· | |
| E | 3773 | hCoV-19/England/BRIS-130297/2020\|EPI_ISL_492614\|20 | ···················· | ···················· | ···················· | ···················· | ···················· | ···················· | |
| E | 3774 | hCoV-19/England/BRIS-130D0F/2020\|EPI_ISL_492615\|20 | ···················· | ···················· | ···················· | ···················· | ···················· | ···················· | |
| E | 3775 | hCoV-19/England/BRIS-130534/2020\|EPI_ISL_492616\|20 | ···················· | ···················· | ···················· | ···················· | ···················· | ···················· | |
| E | 3776 | hCoV-19/England/BRIS-12FE5B/2020\|EPI_ISL_492617\|20 | ···················· | ···················· | ···················· | ···················· | ···················· | ···················· | |
| E | 3777 | hCoV-19/England/BRIS-130376/2020\|EPI_ISL_492618\|20 | ···················· | ···················· | ···················· | ···················· | ···················· | ···················· | |
| E | 3778 | hCoV-19/England/BRIS-130B8D/2020\|EPI_ISL_492619\|20 | ···················· | ···················· | ···················· | ···················· | ···················· | ···················· | |
| E | 3779 | hCoV-19/England/BRIS-130B05/2020\|EPI_ISL_492620\|20 | ···················· | ···················· | ···················· | ···················· | ···················· | ···················· | |
| E | 3780 | hCoV-19/England/BRIS-1307A7/2020\|EPI_ISL_492621\|20 | ···················· | ···················· | ···················· | ···················· | ···················· | ···················· | |
| E | 3781 | hCoV-19/England/BRIS-1307C5/2020\|EPI_ISL_492622\|20 | ···················· | ···················· | ···················· | ···················· | ···················· | ···················· | |
| E | 3782 | hCoV-19/England/BRIS-130A08/2020\|EPI_ISL_492623\|20 | ···················· | ···················· | ···················· | ···················· | ···················· | ···················· | |
| E | 3783 | hCoV-19/England/BRIS-12FC33/2020\|EPI_ISL_492624\|20 | ···················· | ···················· | ···················· | ···················· | ···················· | ···················· | |
| E | 3784 | hCoV-19/England/BRIS-13009D/2020\|EPI_ISL_492625\|20 | ···················· | ···················· | ···················· | ···················· | ···················· | ···················· | |
| E | 3785 | hCoV-19/England/BRIS-130464/2020\|EPI_ISL_492626\|20 | ···················· | ···················· | ···················· | ···················· | ···················· | ···················· | |
| E | 3786 | hCoV-19/England/BRIS-130CC6/2020\|EPI_ISL_492627\|20 | ···················· | ···················· | ···················· | ···················· | ···················· | ···················· | |
| E | 3787 | hCoV-19/England/BRIS-12FE88/2020\|EPI_ISL_492628\|20 | ···················· | ···················· | ···················· | ···················· | ···················· | ···················· | |
| E | 3788 | hCoV-19/England/BRIS-13091A/2020\|EPI_ISL_492629\|20 | ···················· | ···················· | ···················· | ···················· | ···················· | ···················· | |
| E | 3789 | hCoV-19/England/LOND-124C461/2020\|EPI_ISL_492874\|2 | ···················· | ···················· | ···················· | ···················· | ···················· | ···················· | |
| E | 3790 | hCoV-19/England/LOND-124C2D0/2020\|EPI_ISL_492877\|2 | ···················· | ···················· | ···················· | ···················· | ···················· | ···················· | |
| E | 3791 | hCoV-19/England/LOND-124C416/2020\|EPI_ISL_492879\|2 | ···················· | ···················· | ···················· | ···················· | ···················· | ···················· | |
| E | 3792 | hCoV-19/England/LOND-124C58C/2020\|EPI_ISL_492890\|2 | ···················· | ···················· | ···················· | ···················· | ···················· | ···················· | |
| E | 3793 | hCoV-19/England/WSFT-25C1158/2020\|EPI_ISL_492893\|2 | ···················· | ···················· | ···················· | ···················· | ···················· | ···················· | |
| E | 3794 | hCoV-19/England/LOND-124C5E6/2020\|EPI_ISL_492894\|2 | ···················· | ···················· | ···················· | ···················· | ···················· | ···················· | |
| E | 3795 | hCoV-19/England/WSFT-25C12A0/2020\|EPI_ISL_492895\|2 | ···················· | ···················· | ···················· | ···················· | ···················· | ···················· | |
| E | 3796 | hCoV-19/England/WSFT-25C0E61/2020\|EPI_ISL_492896\|2 | ···················· | ···················· | ···················· | ···················· | ···················· | ···················· | |
| E | 3797 | hCoV-19/England/WSFT-25C1194/2020\|EPI_ISL_492898\|2 | ···················· | ···················· | ···················· | ···················· | ···················· | ···················· | |
| E | 3798 | hCoV-19/England/WSFT-25C110D/2020\|EPI_ISL_492899\|2 | ···················· | ···················· | ···················· | ···················· | ···················· | ···················· | |
| E | 3799 | hCoV-19/Scotland/EDB4438/2020\|EPI_ISL_493900\|2020- | ···················· | ···················· | ···················· | ···················· | ···················· | ···················· | |
| E | 3800 | hCoV-19/Scotland/EDB4439/2020\|EPI_ISL_493901\|2020- | ···················· | ···················· | ···················· | ···················· | ···················· | ···················· | |
| E | 3801 | hCoV-19/Scotland/EDB4441/2020\|EPI_ISL_493902\|2020- | ···················· | ···················· | ···················· | ···················· | ···················· | ···················· | |
| E | 3802 | hCoV-19/Scotland/EDB4443/2020\|EPI_ISL_493903\|2020- | ···················· | ···················· | ···················· | ···················· | ···················· | ···················· | |
| E | 3803 | hCoV-19/Scotland/EDB4444/2020\|EPI_ISL_493904\|2020- | ···················· | ···················· | ···················· | ···················· | ···················· | ···················· | |
| E | 3804 | hCoV-19/Scotland/EDB4550/2020\|EPI_ISL_493905\|2020- | ···················· | ···················· | ···················· | ···················· | ···················· | ···················· | |
| E | 3805 | hCoV-19/Scotland/EDB4561/2020\|EPI_ISL_493906\|2020- | ···················· | ···················· | ···················· | ···················· | ···················· | ···················· | |
| E | 3806 | hCoV-19/Scotland/EDB4601/2020\|EPI_ISL_493907\|2020- | ···················· | ···················· | ···················· | ···················· | ···················· | ···················· | |
| E | 3807 | hCoV-19/Scotland/EDB4602/2020\|EPI_ISL_493908\|2020- | ···················· | ···················· | ···················· | ···················· | ···················· | ···················· | |
| E | 3808 | hCoV-19/Scotland/EDB4606/2020\|EPI_ISL_493909\|2020- | ···················· | ···················· | ···················· | ···················· | ···················· | ···················· | |
| E | 3809 | hCoV-19/Scotland/EDB4655/2020\|EPI_ISL_493910\|2020- | ···················· | ·A·················· | ···················· | ···················· | ···················· | ···················· | |
| E | 3810 | hCoV-19/Scotland/EDB4665/2020\|EPI_ISL_493911\|2020- | ···················· | ·A·················· | ···················· | ···················· | ···················· | ···················· | |
| E | 3811 | hCoV-19/Scotland/EDB4689/2020\|EPI_ISL_493912\|2020- | ···················· | ···················· | ···················· | ···················· | ···················· | ···················· | |
| E | 3812 | hCoV-19/Scotland/EDB4692/2020\|EPI_ISL_493913\|2020- | ···················· | ···················· | ···················· | ···················· | ···················· | ···················· | |
| E | 3813 | hCoV-19/Scotland/EDB4695/2020\|EPI_ISL_493914\|2020- | ···················· | ···················· | ···················· | ···················· | ···················· | ···················· | |
| E | 3814 | hCoV-19/Scotland/EDB4698/2020\|EPI_ISL_493916\|2020- | ···················· | ···················· | ···················· | ···················· | ···················· | ···················· | |
| E | 3815 | hCoV-19/Scotland/EDB4903/2020\|EPI_ISL_493919\|2020- | ···················· | ·A·················· | ···················· | ···················· | ···················· | ···················· | |
| E | 3816 | hCoV-19/Scotland/EDB4905/2020\|EPI_ISL_493920\|2020- | ···················· | ·A·················· | ···················· | ···················· | ···················· | ···················· | |
| E | 3817 | hCoV-19/Scotland/EDB4907/2020\|EPI_ISL_493921\|2020- | ···················· | ·A·················· | ···················· | ···················· | ···················· | ···················· | |
| E | 3818 | hCoV-19/Scotland/EDB4912/2020\|EPI_ISL_493922\|2020- | ···················· | ·A·················· | ···················· | ···················· | ···················· | ···················· | |
| E | 3819 | hCoV-19/Scotland/EDB4974/2020\|EPI_ISL_493926\|2020- | ···················· | ···················· | ···················· | ···················· | ···················· | ···················· | |
| E | 3820 | hCoV-19/Scotland/EDB4984/2020\|EPI_ISL_493927\|2020- | ···················· | ···················· | ···················· | ···················· | ···················· | ···················· | |
| E | 3821 | hCoV-19/Scotland/EDB4985/2020\|EPI_ISL_493928\|2020- | ···················· | ·A·················· | ···················· | ···················· | ···················· | ···················· | |
| E | 3822 | hCoV-19/Scotland/EDB4986/2020\|EPI_ISL_493929\|2020- | ···················· | ·A·················· | ···················· | ···················· | ···················· | ···················· | |
| E | 3823 | hCoV-19/Scotland/EDB4989/2020\|EPI_ISL_493930\|2020- | ···················· | ···················· | ···················· | ···················· | ···················· | ···················· | |
| E | 3824 | hCoV-19/Scotland/EDB5008/2020\|EPI_ISL_493932\|2020- | ···················· | ·A·················· | ···················· | ···················· | ···················· | ···················· | |
| E | 3825 | hCoV-19/Scotland/EDB5012/2020\|EPI_ISL_493933\|2020- | ···················· | ·A·················· | ···················· | ···················· | ···················· | ···················· | |
| E | 3826 | hCoV-19/Scotland/EDB5013/2020\|EPI_ISL_493934\|2020- | ···················· | ···················· | ···················· | ···················· | ···················· | ···················· | |
| E | 3827 | hCoV-19/Scotland/EDB5018/2020\|EPI_ISL_493936\|2020- | ···················· | ···················· | ···················· | ···················· | ···················· | ···················· | |
| E | 3828 | hCoV-19/Scotland/EDB5019/2020\|EPI_ISL_493937\|2020- | ···················· | ·A·················· | ···················· | ···················· | ···················· | ···················· | |
| E | 3829 | hCoV-19/Scotland/EDB5028/2020\|EPI_ISL_493938\|2020- | ···················· | ···················· | ···················· | ···················· | ···················· | ···················· | |
| E | 3830 | hCoV-19/Scotland/EDB5029/2020\|EPI_ISL_493939\|2020- | ···················· | ···················· | ···················· | ···················· | ···················· | ···················· | |
| E | 3831 | hCoV-19/Scotland/EDB5031/2020\|EPI_ISL_493941\|2020- | ···················· | ···················· | ···················· | ···················· | ···················· | ···················· | |
| E | 3832 | hCoV-19/Scotland/EDB5153/2020\|EPI_ISL_493943\|2020- | ···················· | ···················A | ···················· | ···················· | ···················· | ···················· | 1 |

| | | | | | | | | |
|---|---|---|---|---|---|---|---|---|
| E | 3833 hCoV-19/Scotland/EDB5154/2020\|EPI_ISL_493944\|2020- | ···················· | ···················· | ···················· | ···················· | ···················· | ···················· | |
| E | 3834 hCoV-19/Scotland/EDB5170/2020\|EPI_ISL_493945\|2020- | ···················· | ···················· | ···················· | ···················· | ···················· | ···················· | |
| E | 3835 hCoV-19/Scotland/EDB5194/2020\|EPI_ISL_493947\|2020- | ···················· | ···················· | ···················· | ···················· | ···················· | ···················· | |
| E | 3836 hCoV-19/Scotland/EDB5270/2020\|EPI_ISL_493949\|2020- | ···················· | ···················A | ···················· | ···················· | ···················· | ···················· | 1 |
| E | 3837 hCoV-19/Scotland/EDB5289/2020\|EPI_ISL_493951\|2020- | ···················· | ·A·················· | ···················· | ···················· | ···················· | ···················· | |
| E | 3838 hCoV-19/Scotland/EDB5320/2020\|EPI_ISL_493952\|2020- | ···················· | ···················· | ···················· | ···················· | ···················· | ···················· | |
| E | 3839 hCoV-19/Scotland/EDB5336/2020\|EPI_ISL_493956\|2020- | ···················· | ···················· | ···················· | ···················· | ···················· | ···················· | |
| E | 3840 hCoV-19/Scotland/EDB5337/2020\|EPI_ISL_493957\|2020- | ···················· | ···················· | ···················· | ···················· | ···················· | ···················· | |
| E | 3841 hCoV-19/Vietnam/IBT-NIHE-03/2020\|EPI_ISL_479756\|20 | ···················· | ···················· | ···················· | ···················· | ···················· | ···················· | |
| E | 3842 hCoV-19/Vietnam/IBT-NIHE-05/2020\|EPI_ISL_479757\|20 | ···················· | ···················· | ···················· | ···················· | ···················· | ···················· | |
| E | 3843 hCoV-19/Vietnam/IBT-NIHE-16/2020\|EPI_ISL_479758\|20 | ···················· | ···················· | ···················· | ···················· | ···················· | ···················· | |
| E | 3844 hCoV-19/USA/FL-04-UMTL-95QV/2020\|EPI_ISL_479759\|20 | ···················· | ···················· | ···················· | ···················· | ···················· | ···················· | |
| E | 3845 hCoV-19/USA/FL-04-UMTL-96NV/2020\|EPI_ISL_479760\|20 | ···················· | ···················· | ···················· | ···················· | ···················· | ···················· | |
| E | 3846 hCoV-19/USA/FL-04-UMTL-975H/2020\|EPI_ISL_479762\|20 | ···················· | ···················· | ···················· | ···················· | ···················· | ···················· | |
| E | 3847 hCoV-19/USA/FL-04-UMTL-9BWN/2020\|EPI_ISL_479763\|20 | ···················· | ···················· | ···················· | ···················· | ···················· | ···················· | |
| E | 3848 hCoV-19/USA/FL-05-UMTL-9TXV/2020\|EPI_ISL_479764\|20 | ···················· | ···················· | ···················· | ···················· | ···················· | ···················· | |
| E | 3849 hCoV-19/USA/FL-06-UMTL-A388/2020\|EPI_ISL_479765\|20 | ···················· | ···················· | ···················· | ···················· | ···················· | ···················· | |
| E | 3850 hCoV-19/USA/FL-06-UMTL-A3MJ/2020\|EPI_ISL_479766\|20 | ···················· | ···················· | ···················· | ···················· | ···················· | ···················· | |
| E | 3851 hCoV-19/USA/FL-06-UMTL-A5QM/2020\|EPI_ISL_479767\|20 | ···················· | ···················· | ···················· | ···················· | ···················· | ···················· | |
| E | 3852 hCoV-19/USA/FL-06-UMTL-A6U1/2020\|EPI_ISL_479768\|20 | ···················· | ···················· | ···················· | ···················· | ···················· | ···················· | |
| E | 3853 hCoV-19/USA/FL-06-UMTL-A6U2/2020\|EPI_ISL_479769\|20 | ···················· | ···················· | ···················· | ···················· | ···················· | ···················· | |
| E | 3854 hCoV-19/USA/FL-06-UMTL-A6U6/2020\|EPI_ISL_479771\|20 | ···················· | ···················· | ···················· | ···················· | ···················· | ···················· | |
| E | 3855 hCoV-19/USA/FL-06-UMTL-A6U7/2020\|EPI_ISL_479772\|20 | ···················· | ···················· | ···················· | ···················· | ···················· | ···················· | |
| E | 3856 hCoV-19/USA/FL-06-UMTL-A81P/2020\|EPI_ISL_479773\|20 | ···················· | ···················· | ···················· | ···················· | ···················· | ···················· | |
| E | 3857 hCoV-19/USA/FL-06-UMTL-A87U/2020\|EPI_ISL_479774\|20 | ···················· | ···················· | ···················· | ···················· | ···················· | ···················· | |
| E | 3858 hCoV-19/Senegal/659/2020\|EPI_ISL_480554\|2020-03-21 | ···················· | ···················· | ···················· | ···················· | ···················· | ···················· | |
| E | 3859 hCoV-19/Senegal/1351/2020\|EPI_ISL_480556\|2020-03-2 | ···················· | ···················· | ···················· | ···················· | ···················· | ···················· | |
| E | 3864 hCoV-19/Australia/VIC2133/2020\|EPI_ISL_480731\|2020 | ···················· | ···················· | ···················· | ···················· | NNN················· | ···················· | 1 |
| E | 3866 hCoV-19/Australia/VIC2135/2020\|EPI_ISL_480733\|2020 | ···················· | ···················· | ···················· | ···················· | ···················· | ···················· | |
| E | 3867 hCoV-19/Australia/VIC2125/2020\|EPI_ISL_480734\|2020 | ···················· | ···················· | ···················· | ···················· | ···················· | ···················· | |
| E | 3869 hCoV-19/Australia/VIC2127/2020\|EPI_ISL_480736\|2020 | ···················· | ···················· | ···················· | ···················· | ···················· | ···················· | |
| E | 3871 hCoV-19/Australia/VIC2137/2020\|EPI_ISL_480738\|2020 | ···················· | ···················· | ···················· | ···················· | ···················· | ···················· | |
| E | 3874 hCoV-19/Australia/VIC2140/2020\|EPI_ISL_480741\|2020 | ···················· | ·A·················· | ···················· | ···················· | ···················· | ··A················· | |
| E | 3876 hCoV-19/Australia/VIC2142/2020\|EPI_ISL_480743\|2020 | ···················· | ···················· | ···················· | ···················· | NNN················· | ···················· | 1 |
| E | 3877 hCoV-19/Australia/VIC2143/2020\|EPI_ISL_480744\|2020 | ···················· | ···················· | ···················· | ···················· | ···················· | ···················· | |
| E | 3880 hCoV-19/Australia/VIC2053/2020\|EPI_ISL_480749\|2020 | ···················· | ···················· | ···················· | ···················· | ···················· | ···················· | |
| E | 3883 hCoV-19/England/BRIS-12D2E8/2020\|EPI_ISL_481861\|20 | ···················· | ···················· | ···················· | ···················· | ···················· | ···················· | |
| E | 3884 hCoV-19/England/BRIS-12D619/2020\|EPI_ISL_481862\|20 | ···················· | ···················· | ···················· | ···················· | ···················· | ···················· | |
| E | 3885 hCoV-19/England/BRIS-12D97A/2020\|EPI_ISL_481863\|20 | ···················· | ···················· | ···················· | ···················· | ···················· | ···················· | |
| E | 3886 hCoV-19/England/BRIS-12DA68/2020\|EPI_ISL_481865\|20 | ···················· | ···················· | ···················· | ···················· | ···················· | ···················· | |
| E | 3887 hCoV-19/England/BRIS-12D4E2/2020\|EPI_ISL_481866\|20 | ···················· | ···················· | ···················· | ···················· | ···················· | ···················· | |
| E | 3888 hCoV-19/England/BRIS-12CD33/2020\|EPI_ISL_481867\|20 | ···················· | ···················· | ···················· | ···················· | ···················· | ···················· | |
| E | 3889 hCoV-19/England/BRIS-12D93E/2020\|EPI_ISL_481868\|20 | ···················· | ···················· | ···················· | ···················· | ···················· | ···················· | |
| E | 3890 hCoV-19/England/BRIS-12CB2A/2020\|EPI_ISL_481869\|20 | ···················· | ···················· | ···················· | ···················· | ···················· | ···················· | |
| E | 3891 hCoV-19/England/BRIS-12D5A3/2020\|EPI_ISL_481870\|20 | ···················· | ···················· | ···················· | ···················· | ···················· | ···················· | |
| E | 3892 hCoV-19/England/BRIS-12D8C8/2020\|EPI_ISL_481871\|20 | ···················· | ···················· | ···················· | ···················· | ···················· | ···················· | |
| E | 3893 hCoV-19/England/BRIS-12D46A/2020\|EPI_ISL_481873\|20 | ···················· | ···················· | ···················· | ···················· | ···················· | ···················· | |
| E | 3894 hCoV-19/England/BRIS-12D34F/2020\|EPI_ISL_481874\|20 | ···················· | ···················· | ···················· | ···················· | ···················· | ···················· | |
| E | 3895 hCoV-19/England/BRIS-12D118/2020\|EPI_ISL_481876\|20 | ···················· | ···················· | ···················· | ···················· | ···················· | ···················· | |
| E | 3896 hCoV-19/England/BRIS-12D488/2020\|EPI_ISL_481877\|20 | ···················· | ···················· | ···················· | ···················· | ···················· | ···················· | |
| E | 3897 hCoV-19/England/BRIS-12D3F4/2020\|EPI_ISL_481878\|20 | ···················· | ···················· | ···················· | ···················· | ···················· | ···················· | |
| E | 3898 hCoV-19/England/BRIS-12DBB0/2020\|EPI_ISL_481879\|20 | ···················· | ···················· | ···················· | ···················· | ···················· | ···················· | |
| E | 3899 hCoV-19/England/BRIS-12D1CD/2020\|EPI_ISL_481882\|20 | ···················· | ···················· | ···················· | ···················· | ···················· | ···················· | |
| E | 3900 hCoV-19/England/BRIS-12CCEB/2020\|EPI_ISL_481883\|20 | ···················· | ···················· | ···················· | ···················· | ···················· | ···················· | |
| E | 3901 hCoV-19/England/BRIS-12CA96/2020\|EPI_ISL_481884\|20 | ···················· | ···················· | ···················· | ···················· | ···················· | ···················· | |
| E | 3902 hCoV-19/England/BRIS-12CE21/2020\|EPI_ISL_481886\|20 | ···················· | ···················· | ···················· | ···················· | ···················· | ···················· | |
| E | 3903 hCoV-19/England/BRIS-12CAF0/2020\|EPI_ISL_481887\|20 | ···················· | ···················· | ···················· | ···················· | ···················· | ···················· | |
| E | 3904 hCoV-19/England/BRIS-12D840/2020\|EPI_ISL_481888\|20 | ···················· | ···················· | ···················· | ···················· | ···················· | ···················· | |
| E | 3905 hCoV-19/England/BRIS-12D725/2020\|EPI_ISL_481890\|20 | ···················· | ···················· | ···················· | ···················· | ···················· | ···················· | |
| E | 3906 hCoV-19/England/BRIS-12D998/2020\|EPI_ISL_481891\|20 | ···················· | ···················· | ···················· | ···················· | ···················· | ···················· | |
| E | 3907 hCoV-19/England/BRIS-12D242/2020\|EPI_ISL_481893\|20 | ···················· | ·A·················· | ···················· | ···················· | ···················· | ···················· | |
| E | 3908 hCoV-19/England/BRIS-12D6BF/2020\|EPI_ISL_481894\|20 | ···················· | ···················· | ···················· | ···················· | ···················· | ···················· | |
| E | 3909 hCoV-19/England/BRIS-12D303/2020\|EPI_ISL_481895\|20 | ···················· | ···················· | ···················· | ···················· | ···················· | ···················· | |
| E | 3910 hCoV-19/England/BRIS-12DB56/2020\|EPI_ISL_481898\|20 | ···················· | ···················· | ···················· | ···················· | ···················· | ···················· | |
| E | 3911 hCoV-19/England/BRIS-12D4C4/2020\|EPI_ISL_481899\|20 | ···················· | ···················· | ···················· | ···················· | ···················· | ···················· | |
| E | 3912 hCoV-19/England/GSTT-265DDA7/2020\|EPI_ISL_484104\|2 | ···················· | ···················· | ···················· | ···················· | ···················· | ···················· | |
| E | 3913 hCoV-19/England/GSTT-265DE0E/2020\|EPI_ISL_484110\|2 | ···················· | ···················· | ···················· | ···················· | ···················· | ···················· | |
| E | 3914 hCoV-19/England/GSTT-265DE59/2020\|EPI_ISL_484115\|2 | ···················· | ···················· | ···················· | ···················· | ···················· | ···················· | |
| E | 3915 hCoV-19/England/GSTT-265DE77/2020\|EPI_ISL_484117\|2 | ···················· | ···················· | ···················· | ···················· | ···················· | ···················· | |
| E | 3916 hCoV-19/England/GSTT-265DF29/2020\|EPI_ISL_484125\|2 | ···················· | ···················· | ···················· | ···················· | ···················· | ···················· | |
| E | 3917 hCoV-19/England/GSTT-265DF38/2020\|EPI_ISL_484126\|2 | ···················· | ···················· | ···················· | ···················· | ···················· | ···················· | |
| E | 3918 hCoV-19/England/GSTT-265DF65/2020\|EPI_ISL_484129\|2 | ···················· | ···················· | ···················· | ···················· | ···················· | ···················· | |
| E | 3919 hCoV-19/England/GSTT-265DF83/2020\|EPI_ISL_484131\|2 | ···················· | ···················· | ···················· | ···················· | ···················· | ···················· | |
| E | 3920 hCoV-19/England/GSTT-265E1F5/2020\|EPI_ISL_484169\|2 | ···················· | ···················· | ···················· | ···················· | ···················· | ···················· | |
| E | 3921 hCoV-19/Poland/IHG_PAS_4_273/2020\|EPI_ISL_485400\|2 | ···················· | ···················· | ···················· | ···················· | ···················· | ···················· | |
| E | 3922 hCoV-19/Bahrain/Hu-1/2020\|EPI_ISL_485401\|2020-06-2 | ···················· | ···················· | ···················· | ···················· | ···················· | ···················· | |
| E | 3926 hCoV-19/Portugal/IGC3860/2020\|EPI_ISL_491211\|2020- | ···················· | ···················· | ···················· | ···················· | ···················· | ···················· | |
| E | 3930 hCoV-19/Portugal/IGC4024/2020\|EPI_ISL_491224\|2020- | ···················· | ···················· | ···················· | ···················· | ···················· | ···················· | |
| E | 3931 hCoV-19/Portugal/IGC4033/2020\|EPI_ISL_491225\|2020- | ···················· | ···················· | ···················· | ···················· | ···················· | ···················· | |
| E | 3932 hCoV-19/Portugal/IGC4089/2020\|EPI_ISL_491226\|2020- | ···················· | ···················· | ···················· | ···················· | ···················· | ···················· | |

| | | | | | | | | |
|---|---|---|---|---|---|---|---|---|
| E | 3933 | hCoV-19/Portugal/IGC4092/2020\|EPI_ISL_491227\|2020- | ···················· | ···················· | ····················· | ···················· | ···················· | ···················· |
| E | 3935 | hCoV-19/Portugal/IGC4096/2020\|EPI_ISL_491229\|2020- | ···················· | ···················· | ····················· | ···················· | ···················· | ···················· |
| E | 3936 | hCoV-19/Portugal/IGC4102/2020\|EPI_ISL_491230\|2020- | ···················· | ···················· | ····················· | ···················· | ···················· | ···················· |
| E | 3937 | hCoV-19/Portugal/IGC4103/2020\|EPI_ISL_491231\|2020- | ···················· | ···················· | ····················· | ···················· | ···················· | ···················· |
| E | 3942 | hCoV-19/Portugal/IGC4234/2020\|EPI_ISL_491238\|2020- | ···················· | ···················· | ····················· | ···················· | ···················· | ···················· |
| E | 3946 | hCoV-19/Portugal/IGC4251/2020\|EPI_ISL_491242\|2020- | ···················· | ···················· | ····················· | ···················· | ···················· | ···················· |
| E | 3948 | hCoV-19/Portugal/IGC4391/2020\|EPI_ISL_491244\|2020- | ···················· | ···················· | ····················· | ···················· | ···················· | ···················· |
| E | 3950 | hCoV-19/Portugal/IGC4395/2020\|EPI_ISL_491246\|2020- | ···················· | ···················· | ····················· | ···················· | ···················· | ···················· |
| E | 3951 | hCoV-19/Portugal/IGC4403/2020\|EPI_ISL_491248\|2020- | ···················· | ···················· | ····················· | ···················· | ···················· | ···················· |
| E | 3953 | hCoV-19/Portugal/IGC4425/2020\|EPI_ISL_491250\|2020- | ···················· | ···················· | ····················· | ···················· | ···················· | ···················· |
| E | 3955 | hCoV-19/Portugal/IGC4435/2020\|EPI_ISL_491252\|2020- | ···················· | ···················· | ····················· | ···················· | ···················· | ···················· |
| E | 3956 | hCoV-19/Portugal/IGC4466/2020\|EPI_ISL_491253\|2020- | ···················· | ···················· | ····················· | ···················· | ···················· | ···················· |
| E | 3957 | hCoV-19/Portugal/IGC4467/2020\|EPI_ISL_491254\|2020- | ···················· | ···················· | ····················· | ···················· | ···················· | ···················· |
| E | 3961 | hCoV-19/Portugal/IGC4478/2020\|EPI_ISL_491258\|2020- | ···················· | ···················· | ····················· | ···················· | ···················· | ···················· |
| E | 3962 | hCoV-19/Portugal/IGC4479/2020\|EPI_ISL_491259\|2020- | ···················· | ···················· | ····················· | ···················· | ···················· | ···················· |
| E | 3963 | hCoV-19/Portugal/IGC4489/2020\|EPI_ISL_491260\|2020- | ···················· | ···················· | ····················· | ···················· | ···················· | ···················· |
| E | 3965 | hCoV-19/Portugal/IGC4549/2020\|EPI_ISL_491262\|2020- | ···················· | ···················· | ····················· | ···················· | ···················· | ···················· |
| E | 3966 | hCoV-19/Portugal/IGC4560/2020\|EPI_ISL_491263\|2020- | ···················· | ···················· | ····················· | ···················· | ···················· | ···················· |
| E | 3967 | hCoV-19/Portugal/R1171/2020\|EPI_ISL_491268\|2020-06 | ···················· | ···················· | ····················· | ···················· | ···················· | ···················· |
| E | 3968 | hCoV-19/Portugal/R1172/2020\|EPI_ISL_491269\|2020-06 | ···················· | ···················· | ····················· | ···················· | ···················· | ···················· |
| E | 3971 | hCoV-19/Portugal/R1291/2020\|EPI_ISL_491273\|2020-06 | ···················· | ···················· | ····················· | ···················· | ···················· | ···················· |
| E | 3973 | hCoV-19/Portugal/R1320/2020\|EPI_ISL_491275\|2020-06 | ···················· | ···················· | ····················· | ···················· | ···················· | ···················· |
| E | 3977 | hCoV-19/Portugal/R998/2020\|EPI_ISL_491297\|2020-06- | ···················· | ···················· | ····················· | ···················· | ···················· | ···················· |
| E | 3978 | hCoV-19/USA/AZ-UW-655/2020\|EPI_ISL_491299\|2020-06- | ···················· | ···················· | ····················· | ···················· | ···················· | ···················· |
| E | 3979 | hCoV-19/USA/WI-UW-735/2020\|EPI_ISL_491410\|2020-07- | ···················· | ···················· | ····················· | ···················· | ···················· | ···················· |
| E | 3980 | hCoV-19/USA/WI-UW-743/2020\|EPI_ISL_491416\|2020-06- | ···················· | ···················· | ····················· | ···················· | ···················· | ···················· |
| E | 3981 | hCoV-19/USA/WI-UW-744/2020\|EPI_ISL_491417\|2020-06- | ···················· | ···················· | ····················· | ···················· | ···················· | ···················· |
| E | 3982 | hCoV-19/USA/WI-UW-745/2020\|EPI_ISL_491418\|2020-07- | ···················· | ···················· | ····················· | ···················· | ···················· | ···················· |
| E | 3983 | hCoV-19/England/BRIS-1301D6/2020\|EPI_ISL_492510\|20 | ···················· | ···················· | ····················· | ···················· | ···················· | ···················· |
| E | 3984 | hCoV-19/England/BRIS-1302F1/2020\|EPI_ISL_492511\|20 | ···················· | ···················· | ····················· | ···················· | ···················· | ···················· |
| E | 3985 | hCoV-19/England/BRIS-12FE4C/2020\|EPI_ISL_492513\|20 | ···················· | ···················· | ····················· | ···················· | ···················· | ···················· |
| E | 3986 | hCoV-19/England/BRIS-130B23/2020\|EPI_ISL_492515\|20 | ···················· | ···················· | ····················· | ···················· | ···················· | ···················· |
| E | 3987 | hCoV-19/England/BRIS-12FD30/2020\|EPI_ISL_492516\|20 | ···················· | ···················· | ····················· | ···················· | ···················· | ···················· |
| E | 3988 | hCoV-19/England/BRIS-12FD03/2020\|EPI_ISL_492517\|20 | ···················· | ···················· | ····················· | ···················· | ···················· | ···················· |
| E | 3989 | hCoV-19/England/BRIS-12FD12/2020\|EPI_ISL_492518\|20 | ···················· | ···················· | ····················· | ···················· | ···················· | ···················· |
| E | 3990 | hCoV-19/England/BRIS-12FF67/2020\|EPI_ISL_492519\|20 | ···················· | ···················· | ····················· | ···················· | ···················· | ···················· |
| E | 3991 | hCoV-19/England/BRIS-1301B8/2020\|EPI_ISL_492520\|20 | ···················· | ···················· | ····················· | ···················· | ···················· | ···················· |
| E | 3992 | hCoV-19/England/BRIS-130570/2020\|EPI_ISL_492521\|20 | ···················· | ···················· | ····················· | ···················· | ···················· | ···················· |
| E | 3993 | hCoV-19/England/BRIS-12FF3A/2020\|EPI_ISL_492522\|20 | ···················· | ···················· | ····················· | ···················· | ···················· | ···················· |
| E | 3994 | hCoV-19/England/BRIS-130C4E/2020\|EPI_ISL_492523\|20 | ···················· | ···················· | ····················· | ···················· | ···················· | ···················· |
| E | 3995 | hCoV-19/England/BRIS-1305AD/2020\|EPI_ISL_492525\|20 | ···················· | ···················· | ····················· | ···················· | ···················· | ···················· |
| E | 3996 | hCoV-19/England/BRIS-1301C7/2020\|EPI_ISL_492527\|20 | ···················· | ···················· | ····················· | ···················· | ···················· | ···················· |
| E | 3997 | hCoV-19/England/BRIS-13066E/2020\|EPI_ISL_492528\|20 | ···················· | ···················· | ····················· | ···················· | ···················· | ···················· |
| E | 3998 | hCoV-19/England/BRIS-13075C/2020\|EPI_ISL_492529\|20 | ···················· | ···················· | ····················· | ···················· | ···················· | ···················· |
| E | 3999 | hCoV-19/England/BRIS-1305F8/2020\|EPI_ISL_492530\|20 | ···················· | ···················· | ····················· | ···················· | ···················· | ···················· |
| E | 4000 | hCoV-19/England/BRIS-13077A/2020\|EPI_ISL_492531\|20 | ···················· | ···················· | ····················· | ···················· | ···················· | ···················· |
| E | 4001 | hCoV-19/England/BRIS-1304DD/2020\|EPI_ISL_492532\|20 | ···················· | ···················· | ····················· | ···················· | ···················· | ···················· |
| E | 4002 | hCoV-19/England/BRIS-12FDB8/2020\|EPI_ISL_492533\|20 | ···················· | ···················· | ····················· | ···················· | ···················· | ···················· |
| E | 4003 | hCoV-19/England/BRIS-12FF0D/2020\|EPI_ISL_492534\|20 | ···················· | ···················· | ····················· | ···················· | ···················· | ···················· |
| E | 4004 | hCoV-19/England/BRIS-130437/2020\|EPI_ISL_492535\|20 | ···················· | ···················· | ····················· | ···················· | ···················· | ···················· |
| E | 4005 | hCoV-19/England/BRIS-130BC9/2020\|EPI_ISL_492536\|20 | ···················· | ···················· | ····················· | ···················· | ···················· | ···················· |
| E | 4006 | hCoV-19/England/BRIS-13067D/2020\|EPI_ISL_492537\|20 | ···················· | ···················· | ····················· | ···················· | ···················· | ···················· |
| E | 4007 | hCoV-19/England/BRIS-130103/2020\|EPI_ISL_492538\|20 | ···················· | ···················· | ····················· | ···················· | ···················· | ···················· |
| E | 4008 | hCoV-19/England/BRIS-13090B/2020\|EPI_ISL_492539\|20 | ···················· | ···················· | ····················· | ···················· | ···················· | ···················· |
| E | 4009 | hCoV-19/England/BRIS-13058F/2020\|EPI_ISL_492540\|20 | ···················· | ···················· | ····················· | ···················· | ···················· | ···················· |
| E | 4010 | hCoV-19/England/BRIS-130288/2020\|EPI_ISL_492541\|20 | ···················· | ···················· | ····················· | ···················· | ···················· | ···················· |
| E | 4011 | hCoV-19/England/BRIS-130446/2020\|EPI_ISL_492543\|20 | ···················· | ···················· | ····················· | ···················· | ···················· | ···················· |
| E | 4012 | hCoV-19/England/BRIS-130B50/2020\|EPI_ISL_492544\|20 | ···················· | ···················· | ····················· | ···················· | ···················· | ···················· |
| E | 4013 | hCoV-19/England/BRIS-13007F/2020\|EPI_ISL_492545\|20 | ···················· | ···················· | ····················· | ···················· | ···················· | ···················· |
| E | 4014 | hCoV-19/England/BRIS-130B14/2020\|EPI_ISL_492547\|20 | ···················· | ···················· | ····················· | ···················· | ···················· | ···················· |
| E | 4015 | hCoV-19/England/BRIS-12FCF7/2020\|EPI_ISL_492548\|20 | ···················· | ···················· | ····················· | ···················· | ···················· | ···················· |
| E | 4016 | hCoV-19/England/BRIS-130798/2020\|EPI_ISL_492549\|20 | ···················· | ···················· | ····················· | ···················· | ···················· | ···················· |
| E | 4017 | hCoV-19/England/BRIS-12FF94/2020\|EPI_ISL_492550\|20 | ···················· | ···················· | ····················· | ···················· | ···················· | ···················· |
| E | 4018 | hCoV-19/England/BRIS-1307E3/2020\|EPI_ISL_492551\|20 | ···················· | ···················· | ····················· | ···················· | ···················· | ···················· |
| E | 4019 | hCoV-19/England/BRIS-12FD21/2020\|EPI_ISL_492552\|20 | ···················· | ···················· | ····················· | ···················· | ···················· | ···················· |
| E | 4020 | hCoV-19/England/BRIS-1306E6/2020\|EPI_ISL_492553\|20 | ···················· | ···················· | ····················· | ···················· | ···················· | ···················· |
| E | 4021 | hCoV-19/England/BRIS-13073E/2020\|EPI_ISL_492554\|20 | ···················· | ···················· | ····················· | ···················· | ···················· | ···················· |
| E | 4022 | hCoV-19/England/BRIS-1303A3/2020\|EPI_ISL_492556\|20 | ···················· | ···················· | ····················· | ···················· | ···················· | ···················· |
| E | 4023 | hCoV-19/England/BRIS-130B7E/2020\|EPI_ISL_492557\|20 | ···················· | ···················· | ····················· | ···················· | ···················· | ···················· |
| E | 4024 | hCoV-19/England/BRIS-130613/2020\|EPI_ISL_492558\|20 | ···················· | ···················· | ····················· | ···················· | ···················· | ···················· |
| E | 4025 | hCoV-19/England/BRIS-1304A0/2020\|EPI_ISL_492559\|20 | ···················· | ···················· | ····················· | ···················· | ···················· | ···················· |
| E | 4026 | hCoV-19/England/BRIS-13017C/2020\|EPI_ISL_492560\|20 | ···················· | ···················· | ····················· | ···················· | ···················· | ···················· |
| E | 4027 | hCoV-19/England/BRIS-1308C2/2020\|EPI_ISL_492561\|20 | ···················· | ·············A······ | ····················· | ···················· | ···················· | ···················· |
| E | 4028 | hCoV-19/England/BRIS-13016D/2020\|EPI_ISL_492562\|20 | ···················· | ···················· | ····················· | ···················· | ···················· | ···················· |
| E | 4029 | hCoV-19/England/BRIS-12FF58/2020\|EPI_ISL_492564\|20 | ···················· | ···················· | ····················· | ···················· | ···················· | ···················· |
| E | 4030 | hCoV-19/England/BRIS-12FF2B/2020\|EPI_ISL_492565\|20 | ···················· | ···················· | ····················· | ···················· | ···················· | ···················· |
| E | 4031 | hCoV-19/England/BRIS-130428/2020\|EPI_ISL_492566\|20 | ···················· | ···················· | ····················· | ···················· | ···················· | ···················· |
| E | 4032 | hCoV-19/England/BRIS-130965/2020\|EPI_ISL_492567\|20 | ···················· | ···················· | ····················· | ···················· | ···················· | ···················· |
| E | 4033 | hCoV-19/England/BRIS-130A53/2020\|EPI_ISL_492569\|20 | ···················· | ···················· | ····················· | ···················· | ···················· | ···················· |
| E | 4034 | hCoV-19/England/BRIS-130D3C/2020\|EPI_ISL_492570\|20 | ···················· | ···················· | ····················· | ···················· | ···················· | ···················· |

| | | | | | | | | |
|---|---|---|---|---|---|---|---|---|
| E | 4035 hCoV-19/England/BRIS-13083B/2020\|EPI_ISL_492571\|20 | .................... | .................... | .................... | .................... | .................... | .................... | |
| E | 4036 hCoV-19/England/BRIS-12FBFA/2020\|EPI_ISL_492572\|20 | .................... | .................... | .................... | .................... | .................... | .................... | |
| E | 4037 hCoV-19/England/BRIS-13024C/2020\|EPI_ISL_492573\|20 | .................... | .................... | .................... | .................... | .................... | .................... | |
| E | 4038 hCoV-19/England/BRIS-1304BF/2020\|EPI_ISL_492574\|20 | .................... | .................... | .................... | .................... | .................... | .................... | |
| E | 4039 hCoV-19/England/BRIS-1309FC/2020\|EPI_ISL_492575\|20 | .................... | .................... | .................... | .................... | .................... | .................... | |
| E | 4040 hCoV-19/England/BRIS-13031C/2020\|EPI_ISL_492576\|20 | .................... | .................... | .................... | .................... | .................... | .................... | |
| E | 4041 hCoV-19/England/BRIS-1308FF/2020\|EPI_ISL_492577\|20 | .................... | .................... | .................... | .................... | .................... | .................... | |
| E | 4042 hCoV-19/England/BRIS-13014F/2020\|EPI_ISL_492578\|20 | .................... | .................... | .................... | .................... | .................... | .................... | |
| E | 4043 hCoV-19/England/BRIS-130112/2020\|EPI_ISL_492579\|20 | .................... | .................... | .................... | .................... | .................... | .................... | |
| E | 4044 hCoV-19/England/BRIS-12FC15/2020\|EPI_ISL_492580\|20 | .................... | .................... | .................... | .................... | .................... | .................... | |
| E | 4045 hCoV-19/England/BRIS-12FDE5/2020\|EPI_ISL_492581\|20 | .................... | .................... | .................... | .................... | .................... | .................... | |
| E | 4046 hCoV-19/England/BRIS-130358/2020\|EPI_ISL_492582\|20 | .................... | .................... | .................... | .................... | .................... | .................... | |
| E | 4047 hCoV-19/England/BRIS-12FEB5/2020\|EPI_ISL_492583\|20 | .................... | .................... | .................... | .................... | .................... | .................... | |
| E | 4048 hCoV-19/England/BRIS-12FE2E/2020\|EPI_ISL_492584\|20 | .................... | .................... | .................... | .................... | .................... | .................... | |
| E | 4049 hCoV-19/England/BRIS-1304CE/2020\|EPI_ISL_492586\|20 | .................... | .................... | .................... | .................... | .................... | .................... | |
| E | 4050 hCoV-19/England/BRIS-13080E/2020\|EPI_ISL_492587\|20 | .................... | .................... | .................... | .................... | .................... | .................... | |
| E | 4051 hCoV-19/England/BRIS-130631/2020\|EPI_ISL_492589\|20 | .................... | .................... | .................... | .................... | .................... | .................... | |
| E | 4052 hCoV-19/England/BRIS-130C3F/2020\|EPI_ISL_492591\|20 | .................... | .................... | .................... | .................... | .................... | .................... | |
| E | 4053 hCoV-19/England/BRIS-130BAB/2020\|EPI_ISL_492592\|20 | .................... | .................... | .................... | .................... | .................... | .................... | |
| E | 4054 hCoV-19/England/BRIS-130859/2020\|EPI_ISL_492595\|20 | .................... | .................... | .................... | A................... | .................... | .................... | |
| E | 4055 hCoV-19/England/BRIS-13081D/2020\|EPI_ISL_492597\|20 | .................... | .................... | .................... | .................... | .................... | .................... | |
| E | 4056 hCoV-19/England/BRIS-130640/2020\|EPI_ISL_492598\|20 | .................... | .................... | .................... | .................... | .................... | .................... | |
| E | 4057 hCoV-19/England/BRIS-12FCBB/2020\|EPI_ISL_492599\|20 | .................... | .................... | .................... | .................... | .................... | .................... | |
| E | 4058 hCoV-19/England/BRIS-130A62/2020\|EPI_ISL_492710\|20 | .................... | .................... | .................... | .................... | .................... | .................... | |
| E | 4059 hCoV-19/England/BRIS-130015/2020\|EPI_ISL_492711\|20 | .................... | .................... | .................... | .................... | .................... | .................... | |
| E | 4060 hCoV-19/England/BRIS-130419/2020\|EPI_ISL_492712\|20 | .................... | .................... | .................... | .................... | .................... | .................... | |
| E | 4061 hCoV-19/England/BRIS-130024/2020\|EPI_ISL_492713\|20 | .................... | .................... | .................... | .................... | .................... | .................... | |
| E | 4062 hCoV-19/England/BRIS-13074D/2020\|EPI_ISL_492714\|20 | .................... | .................... | .................... | .................... | .................... | .................... | |
| E | 4063 hCoV-19/England/BRIS-12FDF4/2020\|EPI_ISL_492715\|20 | .................... | .................... | .................... | .................... | .................... | .................... | |
| E | 4064 hCoV-19/England/BRIS-1302C4/2020\|EPI_ISL_492716\|20 | .................... | .................... | .................... | .................... | .................... | .................... | |
| E | 4065 hCoV-19/England/BRIS-1305CB/2020\|EPI_ISL_492717\|20 | .................... | .................... | .................... | .................... | .................... | .................... | |
| E | 4066 hCoV-19/England/BRIS-1309DE/2020\|EPI_ISL_492719\|20 | .................... | .................... | .................... | .................... | .................... | .................... | |
| E | 4067 hCoV-19/England/BRIS-12FCAC/2020\|EPI_ISL_492720\|20 | .................... | .................... | .................... | .................... | .................... | .................... | |
| E | 4068 hCoV-19/England/BRIS-12FFD0/2020\|EPI_ISL_492722\|20 | .................... | .................... | .................... | .................... | .................... | .................... | |
| E | 4069 hCoV-19/England/BRIS-12FF49/2020\|EPI_ISL_492723\|20 | .................... | .................... | .................... | .................... | .................... | .................... | |
| E | 4070 hCoV-19/England/BRIS-13030D/2020\|EPI_ISL_492725\|20 | .................... | .............A...... | .................... | .................... | .................... | .................... | |
| E | 4071 hCoV-19/England/BRIS-130ADB/2020\|EPI_ISL_492726\|20 | .................... | .................... | .................... | .................... | .................... | .................... | |
| E | 4072 hCoV-19/England/BRIS-130C11/2020\|EPI_ISL_492727\|20 | .................... | .................... | .................... | .................... | .................... | .................... | |
| E | 4073 hCoV-19/England/BRIS-12FE00/2020\|EPI_ISL_492728\|20 | .................... | .................... | .................... | .................... | .................... | .................... | |
| E | 4074 hCoV-19/England/BRIS-12FFA3/2020\|EPI_ISL_492729\|20 | .................... | .................... | .................... | .................... | .................... | .................... | |
| E | 4075 hCoV-19/England/BRIS-130279/2020\|EPI_ISL_492740\|20 | .................... | .................... | .................... | .................... | .................... | .................... | |
| E | 4076 hCoV-19/England/BRIS-12FDA9/2020\|EPI_ISL_492742\|20 | .................... | .................... | .................... | .................... | .................... | .................... | |
| E | 4077 hCoV-19/England/BRIS-130121/2020\|EPI_ISL_492744\|20 | .................... | .................... | .................... | .................... | .................... | .................... | |
| E | 4078 hCoV-19/England/BRIS-12FD6D/2020\|EPI_ISL_492745\|20 | .................... | .................... | .................... | .................... | .................... | .................... | |
| E | 4080 hCoV-19/Scotland/CVR3275/2020\|EPI_ISL_493844\|2020- | .................... | ...................A | .................... | .................... | .................... | .................... | 1 |
| E | 4083 hCoV-19/Scotland/CVR3296/2020\|EPI_ISL_493853\|2020- | .................... | ...................A | .................... | .................... | .................... | .................... | 1 |
| E | 4084 hCoV-19/Scotland/CVR3301/2020\|EPI_ISL_493857\|2020- | .................... | .................... | .................... | .................... | .................... | .................... | |
| E | 4085 hCoV-19/Scotland/CVR3302/2020\|EPI_ISL_493858\|2020- | .................... | .................... | .................... | .................... | .................... | .................... | |
| E | 4088 hCoV-19/Scotland/CVR3325/2020\|EPI_ISL_493877\|2020- | .................... | .................... | .................... | .................... | .................... | .................... | |
| E | 4089 hCoV-19/Scotland/CVR3327/2020\|EPI_ISL_493878\|2020- | .................... | .................... | .................... | .................... | .................... | .................... | |
| E | 4090 hCoV-19/Scotland/CVR3331/2020\|EPI_ISL_493882\|2020- | .................... | .................... | .................... | .................... | .................... | .................... | |
| E | 4091 hCoV-19/Scotland/CVR3875/2020\|EPI_ISL_493895\|2020- | .................... | ...................A | .................... | .................... | .................... | .................... | 1 |
| E | 4093 hCoV-19/Scotland/EDB3907/2020\|EPI_ISL_493898\|2020- | .................... | .................... | .................... | .................... | .................... | .................... | |
| E | 4094 hCoV-19/Scotland/EDB4437/2020\|EPI_ISL_493899\|2020- | .................... | .A.................. | .................... | .................... | .................... | .................... | |
| E | 4095 hCoV-19/Germany/BY-UHR-1ZK/2020\|EPI_ISL_513298\|202 | .................... | .................... | .................... | .................... | .................... | .................... | |
| E | 4096 hCoV-19/Germany/BY-UHR-2ZK/2020\|EPI_ISL_513299\|202 | .................... | .................... | .................... | .................... | .................... | .................... | |
| E | 4097 hCoV-19/USA/FL-06-UMTL-A8WJ/2020\|EPI_ISL_479775\|20 | .................... | .................... | .................... | .................... | .................... | .................... | |
| E | 4098 hCoV-19/England/NP1/2020\|EPI_ISL_479777\|2020 | .................... | .................... | .................... | .................... | .................... | .................... | |
| E | 4099 hCoV-19/England/NP2/2020\|EPI_ISL_479778\|2020 | .................... | .................... | .................... | .................... | .................... | .................... | |
| E | 4100 hCoV-19/England/NP4/2020\|EPI_ISL_479780\|2020 | .................... | .................... | .................... | .................... | .................... | .................... | |
| E | 4101 hCoV-19/England/NP5/2020\|EPI_ISL_479781\|2020 | .................... | .................... | .................... | .................... | .................... | .................... | |
| E | 4102 hCoV-19/England/NP6/2020\|EPI_ISL_479782\|2020 | .................... | .................... | .................... | .................... | .................... | .................... | |
| E | 4103 hCoV-19/England/NP7/2020\|EPI_ISL_479783\|2020 | .................... | .................... | .................... | .................... | .................... | .................... | |
| E | 4104 hCoV-19/England/NP8/2020\|EPI_ISL_479784\|2020 | .................... | .................... | .................... | .................... | .................... | .................... | |
| E | 4105 hCoV-19/England/P1/2020\|EPI_ISL_479785\|2020 | .................... | .................... | .................... | .................... | .................... | .................... | |
| E | 4106 hCoV-19/England/P2/2020\|EPI_ISL_479786\|2020 | .................... | .................... | .................... | .................... | .................... | .................... | |
| E | 4107 hCoV-19/England/P4/2020\|EPI_ISL_479788\|2020 | .................... | .................... | .................... | .................... | .................... | .................... | |
| E | 4108 hCoV-19/Australia/VIC2089/2020\|EPI_ISL_480701\|2020 | .................... | .................... | .................... | .................... | .................... | .................... | |
| E | 4109 hCoV-19/Australia/VIC2090/2020\|EPI_ISL_480702\|2020 | .................... | .................... | .................... | .................... | .................... | .................... | |
| E | 4110 hCoV-19/Australia/VIC2093/2020\|EPI_ISL_480704\|2020 | .................... | .................... | .................... | .................... | .................... | .................... | |
| E | 4115 hCoV-19/Australia/VIC2102/2020\|EPI_ISL_480710\|2020 | .................... | .................... | .................... | .................... | .................... | .................... | |
| E | 4116 hCoV-19/Australia/VIC2103/2020\|EPI_ISL_480711\|2020 | .................... | .................... | .................... | .................... | .................... | .................... | |
| E | 4117 hCoV-19/Australia/VIC2104/2020\|EPI_ISL_480712\|2020 | .................... | .................... | .................... | .................... | .................... | .................... | |
| E | 4118 hCoV-19/Australia/VIC2105/2020\|EPI_ISL_480713\|2020 | .................... | .................... | .................... | .................... | .................... | .................... | |
| E | 4122 hCoV-19/Australia/VIC2111/2020\|EPI_ISL_480717\|2020 | .................... | .................... | .................... | .................... | NNN................. | .................... | 1 |
| E | 4123 hCoV-19/Australia/VIC2112/2020\|EPI_ISL_480718\|2020 | .................... | .................... | .................... | .................... | .................... | .................... | |
| E | 4124 hCoV-19/Australia/VIC2113/2020\|EPI_ISL_480719\|2020 | .................... | .................... | .................... | .................... | .................... | .................... | |
| E | 4125 hCoV-19/Australia/VIC2116/2020\|EPI_ISL_480721\|2020 | .................... | .................... | .................... | .................... | .................... | .................... | |
| E | 4127 hCoV-19/Australia/VIC2118/2020\|EPI_ISL_480723\|2020 | .................... | .................... | .................... | .................... | .................... | .................... | |
| E | 4131 hCoV-19/Australia/VIC2129/2020\|EPI_ISL_480728\|2020 | .................... | .................... | .................... | .................... | .................... | .................... | |

| | | | | | | | | |
|---|---|---|---|---|---|---|---|---|
| E | 4132 hCoV-19/Australia/VIC2130/2020\|EPI_ISL_480729\|2020 | ···················· | ···················· | ···················· | ···················· | ···················· | ···················· | |
| E | 4133 hCoV-19/Australia/VIC2054/2020\|EPI_ISL_480750\|2020 | ···················· | ···················· | ···················· | ···················· | ···················· | ···················· | |
| E | 4135 hCoV-19/Australia/VIC2065/2020\|EPI_ISL_480755\|2020 | ···················· | ···················· | ···················· | ···················· | ···················· | ···················· | |
| E | 4136 hCoV-19/Australia/VIC2066/2020\|EPI_ISL_480756\|2020 | ···················· | ···················· | ···················· | ···················· | NNN················· | ···················· | 1 |
| E | 4137 hCoV-19/Australia/VIC2067/2020\|EPI_ISL_480757\|2020 | ···················· | ···················· | ···················· | ···················· | ···················· | ···················· | |
| E | 4138 hCoV-19/Australia/VIC2069/2020\|EPI_ISL_480758\|2020 | ···················· | ···················· | ···················· | ···················· | ···················· | ···················· | |
| E | 4139 hCoV-19/Australia/VIC2068/2020\|EPI_ISL_480759\|2020 | ···················· | ···················· | ···················· | ···················· | ···················· | ···················· | |
| E | 4140 hCoV-19/Australia/VIC2160/2020\|EPI_ISL_480760\|2020 | ···················· | ···················· | ···················· | ···················· | ···················· | ···················· | |
| E | 4141 hCoV-19/Australia/VIC2158/2020\|EPI_ISL_480761\|2020 | ···················· | ···················· | ···················· | ···················· | NNN················· | ···················· | 1 |
| E | 4145 hCoV-19/Australia/VIC2151/2020\|EPI_ISL_480765\|2020 | ···················· | ···················· | ···················· | ···················· | ···················· | ···················· | |
| E | 4149 hCoV-19/Australia/VIC2156/2020\|EPI_ISL_480769\|2020 | NNN················· | ···················· | ···················· | ···················· | ···················· | ···················· | 1 |
| E | 4150 hCoV-19/England/HECH-259A46E/2020\|EPI_ISL_484219\|2 | ···················· | ···················· | ···················· | ···················· | ···················· | ···················· | |
| E | 4151 hCoV-19/England/HECH-259A48C/2020\|EPI_ISL_484220\|2 | ···················· | ···················· | ···················· | ···················· | ···················· | ···················· | |
| E | 4152 hCoV-19/England/LCST-24D6BAE/2020\|EPI_ISL_484222\|2 | ···················· | ···················· | ···················· | ···················· | ···················· | ···················· | |
| E | 4153 hCoV-19/England/LCST-24D6BBD/2020\|EPI_ISL_484223\|2 | ···················· | ···················· | ···················· | ···················· | ···················· | ···················· | |
| E | 4154 hCoV-19/England/LCST-24D6C9C/2020\|EPI_ISL_484225\|2 | ···················· | ···················· | ···················· | ···················· | ···················· | ···················· | |
| E | 4155 hCoV-19/England/LCST-24D6CAB/2020\|EPI_ISL_484226\|2 | ···················· | ···················· | ···················· | ···················· | ···················· | ···················· | |
| E | 4156 hCoV-19/England/LCST-24D6CC9/2020\|EPI_ISL_484228\|2 | ···················· | ···················· | ···················· | ···················· | ···················· | ···················· | |
| E | 4157 hCoV-19/England/LCST-24D6CE7/2020\|EPI_ISL_484229\|2 | ···················· | ···················· | ···················· | ···················· | ···················· | ···················· | |
| E | 4158 hCoV-19/England/LCST-24D6D20/2020\|EPI_ISL_484230\|2 | ···················· | ···················· | ···················· | ···················· | ···················· | ···················· | |
| E | 4159 hCoV-19/England/LCST-24D6D3F/2020\|EPI_ISL_484231\|2 | ···················· | ···················· | ···················· | ···················· | ···················· | ···················· | |
| E | 4160 hCoV-19/England/LCST-24D6D4E/2020\|EPI_ISL_484232\|2 | ···················· | ···················· | ···················· | ···················· | ···················· | ···················· | |
| E | 4161 hCoV-19/England/LCST-24D6DB7/2020\|EPI_ISL_484235\|2 | ···················· | ···················· | ···················· | ···················· | ···················· | ···················· | |
| E | 4162 hCoV-19/England/LCST-24D6DE4/2020\|EPI_ISL_484238\|2 | ···················· | ···················· | ···················· | ···················· | ···················· | ···················· | |
| E | 4163 hCoV-19/England/LCST-24D6DF3/2020\|EPI_ISL_484239\|2 | ···················· | ···················· | ···················· | ···················· | ···················· | ···················· | |
| E | 4164 hCoV-19/England/LCST-24D6E3C/2020\|EPI_ISL_484242\|2 | ···················· | ···················· | ···················· | ···················· | ···················· | ···················· | |
| E | 4165 hCoV-19/England/LCST-24D6E69/2020\|EPI_ISL_484244\|2 | ···················· | ···················· | ···················· | ···················· | ···················· | ···················· | |
| E | 4166 hCoV-19/England/LCST-24D6E96/2020\|EPI_ISL_484245\|2 | ···················· | ···················· | ···················· | ···················· | ···················· | ···················· | |
| E | 4167 hCoV-19/England/LCST-24D6ED2/2020\|EPI_ISL_484249\|2 | ···················· | ···················· | ···················· | ···················· | ···················· | ···················· | |
| E | 4168 hCoV-19/England/LCST-24D6F2A/2020\|EPI_ISL_484251\|2 | ···················· | ···················· | ···················· | ···················· | ···················· | ···················· | |
| E | 4169 hCoV-19/England/LIVE-A198E/2020\|EPI_ISL_484254\|202 | ···················· | ···················· | ···················· | ···················· | ···················· | ···················· | |
| E | 4170 hCoV-19/England/LIVE-A19CA/2020\|EPI_ISL_484256\|202 | ···················· | ···················· | ···················· | ···················· | ···················· | ···················· | |
| E | 4171 hCoV-19/England/LIVE-A1B1F/2020\|EPI_ISL_484262\|202 | ···················· | ···················· | ···················· | ···················· | ···················· | ···················· | |
| E | 4172 hCoV-19/England/LIVE-A1B4C/2020\|EPI_ISL_484263\|202 | ···················· | ···················· | ···················· | ···················· | ···················· | ···················· | |
| E | 4173 hCoV-19/England/NORT-296191/2020\|EPI_ISL_484264\|20 | ···················· | ···················· | ···················· | ···················· | ···················· | ···················· | |
| E | 4174 hCoV-19/England/NORT-2961EC/2020\|EPI_ISL_484265\|20 | ···················· | ···················· | ···················· | ···················· | ···················· | ···················· | |
| E | 4175 hCoV-19/England/NORT-2962DA/2020\|EPI_ISL_484267\|20 | ···················· | ···················· | ···················· | ···················· | ···················· | ···················· | |
| E | 4176 hCoV-19/England/NORT-2963F5/2020\|EPI_ISL_484269\|20 | ···················· | ···················· | ···················· | ···················· | ···················· | ···················· | |
| E | 4177 hCoV-19/England/NORT-2968C9/2020\|EPI_ISL_484270\|20 | ···················· | ···················· | ···················· | ···················· | ···················· | ···················· | |
| E | 4178 hCoV-19/England/NORT-296920/2020\|EPI_ISL_484271\|20 | ···················· | ···················· | ···················· | ···················· | ···················· | ···················· | |
| E | 4179 hCoV-19/England/NORT-296AB4/2020\|EPI_ISL_484273\|20 | ···················· | ···················· | ···················· | ···················· | ···················· | ···················· | |
| E | 4180 hCoV-19/England/NORT-296C27/2020\|EPI_ISL_484274\|20 | ···················· | ···················· | ···················· | ···················· | ···················· | ···················· | |
| E | 4181 hCoV-19/England/NORT-296C45/2020\|EPI_ISL_484275\|20 | ···················· | ···················· | ···················· | ···················· | ···················· | ···················· | |
| E | 4182 hCoV-19/England/NORT-296C81/2020\|EPI_ISL_484276\|20 | ···················· | ···················· | ···················· | ···················· | ···················· | ···················· | |
| E | 4183 hCoV-19/England/NORT-296CBE/2020\|EPI_ISL_484277\|20 | ···················· | ···················· | ···················· | ···················· | ···················· | ···················· | |
| E | 4184 hCoV-19/England/NORT-296CFA/2020\|EPI_ISL_484279\|20 | ···················· | ···················· | ···················· | ···················· | ···················· | ···················· | |
| E | 4185 hCoV-19/England/NORT-296D33/2020\|EPI_ISL_484280\|20 | ···················· | ···················· | ···················· | ···················· | ···················· | ···················· | |
| E | 4186 hCoV-19/England/NORT-296FA6/2020\|EPI_ISL_484282\|20 | ···················· | ···················· | ···················· | ···················· | ···················· | ···················· | |
| E | 4187 hCoV-19/England/NORT-297190/2020\|EPI_ISL_484283\|20 | ···················· | ···················· | ···················· | ···················· | ···················· | ···················· | |
| E | 4188 hCoV-19/England/NORT-29C9EE/2020\|EPI_ISL_484288\|20 | ···················· | ···················· | ···················· | ···················· | ···················· | ···················· | |
| E | 4189 hCoV-19/England/NORT-29CACD/2020\|EPI_ISL_484289\|20 | ···················· | ···················· | ···················· | ···················· | ···················· | ···················· | |
| E | 4190 hCoV-19/England/NORT-29CADC/2020\|EPI_ISL_484290\|20 | ···················· | ···················· | ···················· | ···················· | ···················· | ···················· | |
| E | 4191 hCoV-19/England/NORT-29CB24/2020\|EPI_ISL_484292\|20 | ···················· | ···················· | ···················· | ···················· | ···················· | ···················· | |
| E | 4192 hCoV-19/England/NORT-29CBCA/2020\|EPI_ISL_484293\|20 | ···················· | ···················· | ···················· | ···················· | ···················· | ···················· | |
| E | 4193 hCoV-19/England/NORT-29CC6D/2020\|EPI_ISL_484295\|20 | ···················· | ···················· | ···················· | ···················· | ···················· | ···················· | |
| E | 4194 hCoV-19/Russia/CRIE159560/2020\|EPI_ISL_486815\|2020 | ···················· | ···················· | ···················· | ···················· | ···················· | ···················· | |
| E | 4195 hCoV-19/Russia/CRIE159875/2020\|EPI_ISL_486816\|2020 | ···················· | ···················· | ···················· | ···················· | ···················· | ···················· | |
| E | 4196 hCoV-19/Russia/CRIE159932/2020\|EPI_ISL_486817\|2020 | ···················· | ···················· | ···················· | ···················· | ···················· | ···················· | |
| E | 4197 hCoV-19/Russia/CRIE160277/2020\|EPI_ISL_486818\|2020 | ···················· | ···················· | ···················· | ···················· | ···················· | ···················· | |
| E | 4198 hCoV-19/Russia/CRIE160790/2020\|EPI_ISL_486819\|2020 | ···················· | ···················· | ···················· | ···················· | ···················· | ···················· | |
| E | 4199 hCoV-19/Russia/CRIE164298/2020\|EPI_ISL_486822\|2020 | ···················· | ···················· | ···················· | ···················· | ···················· | ···················· | |
| E | 4200 hCoV-19/Russia/CRIE164399/2020\|EPI_ISL_486823\|2020 | ···················· | ···················· | ···················· | ···················· | ···················· | ···················· | |
| E | 4201 hCoV-19/USA/OR-PROV-029/2020\|EPI_ISL_486830\|2020-0 | ···················· | ···················· | ···················· | ···················· | ···················· | ···················· | |
| E | 4202 hCoV-19/USA/OR-PROV-034/2020\|EPI_ISL_486831\|2020-0 | ···················· | ···················· | ···················· | ···················· | ···················· | ···················· | |
| E | 4203 hCoV-19/Romania/ROSV-2/2020\|EPI_ISL_486834\|2020-05 | ···················· | ···················· | ···················· | ···················· | ···················· | ···················· | |
| E | 4204 hCoV-19/India/KA-InStem-NCBS-0087/2020\|EPI_ISL_486 | ···················· | ···················· | ···················· | ···················· | ···················· | ···················· | |
| E | 4205 hCoV-19/India/KA-InStem-NCBS-0088/2020\|EPI_ISL_486 | ···················· | ···················· | ···················· | ···················· | ···················· | ···················· | |
| E | 4206 hCoV-19/India/KA-InStem-NCBS-0089/2020\|EPI_ISL_486 | ···················· | ···················· | ···················· | ···················· | ···················· | ···················· | |
| E | 4207 hCoV-19/India/KA-InStem-NCBS-0090/2020\|EPI_ISL_486 | ···················· | ···················· | ···················· | ···················· | ···················· | ···················· | |
| E | 4208 hCoV-19/India/KA-InStem-NCBS-0092/2020\|EPI_ISL_486 | ···················· | ···················· | ···················· | ···················· | ···················· | ···················· | |
| E | 4209 hCoV-19/India/KA-InStem-NCBS-0097/2020\|EPI_ISL_486 | ···················· | ···················· | ···················· | ···················· | ···················· | ···················· | |
| E | 4210 hCoV-19/Ecuador/USFQ-105/2020\|EPI_ISL_486842\|2020- | ···················· | ···················· | ···················· | ···················· | ···················· | ···················· | |
| E | 4211 hCoV-19/Ecuador/USFQ-108/2020\|EPI_ISL_486843\|2020- | ···················· | ···················· | ···················· | ···················· | ···················· | ···················· | |
| E | 4212 hCoV-19/Ecuador/USFQ-109/2020\|EPI_ISL_486844\|2020- | ···················· | ···················· | ···················· | ···················· | ···················· | ···················· | |
| E | 4213 hCoV-19/Ecuador/USFQ-110/2020\|EPI_ISL_486845\|2020- | ···················· | ···················· | ···················· | ···················· | ···················· | ···················· | |
| E | 4214 hCoV-19/Ecuador/USFQ-111/2020\|EPI_ISL_486846\|2020- | ···················· | ···················· | ···················· | ···················· | ···················· | ···················· | |
| E | 4215 hCoV-19/Ecuador/USFQ-1112/2020\|EPI_ISL_486847\|2020 | ···················· | ···················· | ···················· | ···················· | ···················· | ···················· | |
| E | 4216 hCoV-19/Ecuador/USFQ-113/2020\|EPI_ISL_486848\|2020- | ···················· | ···················· | ···················· | ···················· | ···················· | ···················· | |
| E | 4217 hCoV-19/Ecuador/USFQ-114/2020\|EPI_ISL_486849\|2020- | ···················· | ···················· | ···················· | ···················· | ···················· | ···················· | |
| E | 4218 hCoV-19/Ecuador/USFQ-118/2020\|EPI_ISL_486850\|2020- | ···················· | ···················· | ···················· | ···················· | ···················· | ···················· | |

| | | | | | | | | |
|---|---|---|---|---|---|---|---|---|
| E | 4219 | hCoV-19/Ecuador/USFQ-119/2020\|EPI_ISL_486851\|2020- | .................... | .................... | .................... | .................... | .................... | .................... |
| E | 4220 | hCoV-19/India/UN-UN1/2020\|EPI_ISL_486852\|2020-06-1 | .................... | .................... | .................... | .................... | .................... | .................... |
| E | 4221 | hCoV-19/India/UN-UN2/2020\|EPI_ISL_486853\|2020-06-1 | .................... | .................... | .................... | .................... | .................... | .................... |
| E | 4222 | hCoV-19/Romania/ROSV_12646/2020\|EPI_ISL_486854\|202 | .................... | .................... | .................... | .................... | .................... | .................... |
| E | 4223 | hCoV-19/Romania/ROSV_12287/2020\|EPI_ISL_486856\|202 | .................... | .................... | .................... | .................... | .................... | .................... |
| E | 4224 | hCoV-19/Senegal/6751/2020\|EPI_ISL_486859\|2020-04-2 | .................... | .................... | .................... | .................... | .................... | .................... |
| E | 4225 | hCoV-19/Senegal/9615/2020\|EPI_ISL_486860\|2020-04-2 | .................... | .................... | .................... | .................... | .................... | .................... |
| E | 4226 | hCoV-19/Senegal/9439/2020\|EPI_ISL_486861\|2020-04-2 | .................... | .................... | .................... | .................... | .................... | .................... |
| E | 4227 | hCoV-19/Senegal/9392/2020\|EPI_ISL_486862\|2020-04-2 | .................... | .................... | .................... | .................... | .................... | .................... |
| E | 4228 | hCoV-19/Senegal/9369/2020\|EPI_ISL_486863\|2020-04-2 | .................... | .................... | .................... | .................... | .................... | .................... |
| E | 4229 | hCoV-19/Senegal/9256/2020\|EPI_ISL_486864\|2020-04-2 | .................... | .................... | .................... | .................... | .................... | .................... |
| E | 4230 | hCoV-19/Senegal/6847/2020\|EPI_ISL_486865\|2020-04-0 | .................... | .................... | .................... | .................... | .................... | .................... |
| E | 4231 | hCoV-19/Senegal/9250/2020\|EPI_ISL_486866\|2020-04-2 | .................... | .................... | .................... | .................... | .................... | .................... |
| E | 4232 | hCoV-19/Senegal/6839/2020\|EPI_ISL_486867\|2020-04-2 | .................... | .................... | .................... | .................... | .................... | .................... |
| E | 4233 | hCoV-19/Senegal/6837/2020\|EPI_ISL_486868\|2020-04-2 | .................... | .................... | .................... | .................... | .................... | .................... |
| E | 4234 | hCoV-19/Senegal/6597/2020\|EPI_ISL_486870\|2020-04-2 | .................... | .................... | .................... | .................... | .................... | .................... |
| E | 4235 | hCoV-19/Senegal/6481/2020\|EPI_ISL_486871\|2020-04-2 | .................... | .................... | .................... | .................... | .................... | .................... |
| E | 4236 | hCoV-19/Senegal/6430/2020\|EPI_ISL_486872\|2020-04-1 | .................... | .................... | .................... | .................... | .................... | .................... |
| E | 4237 | hCoV-19/Senegal/6427/2020\|EPI_ISL_486873\|2020-04-2 | .................... | .................... | .................... | .................... | .................... | .................... |
| E | 4238 | hCoV-19/Spain/PV-HUB-50399382/2020\|EPI_ISL_486876\| | .................... | .................... | .................... | .................... | .................... | .................... |
| E | 4239 | hCoV-19/India/KA-nimh-3975/2020\|EPI_ISL_486881\|202 | .................... | .................... | .................... | .................... | .................... | .................... |
| E | 4240 | hCoV-19/Bahrain/02/2020\|EPI_ISL_486887\|2020-06-23 | .................... | .................... | .................... | .................... | .................... | .................... |
| E | 4241 | hCoV-19/Bahrain/03/2020\|EPI_ISL_486888\|2020-06-23 | .................... | .................... | .................... | .................... | .................... | .................... |
| E | 4242 | hCoV-19/Bahrain/04/2020\|EPI_ISL_486889\|2020-06-23 | .................... | .................... | .................... | .................... | .................... | .................... |
| E | 4243 | hCoV-19/Japan/TK-YE-615419/2020\|EPI_ISL_486897\|202 | .................... | .................... | .................... | .................... | .................... | .................... |
| E | 4244 | hCoV-19/Japan/TK-YE615484/2020\|EPI_ISL_486898\|2020 | .................... | .................... | .................... | .................... | .................... | .................... |
| E | 4245 | hCoV-19/Japan/TK-YE615664/2020\|EPI_ISL_486899\|2020 | .................... | .................... | .................... | .................... | .................... | .................... |
| E | 4246 | hCoV-19/England/NORT-29247E/2020\|EPI_ISL_489070\|20 | .................... | .................... | .................... | .................... | .................... | .................... |
| E | 4247 | hCoV-19/England/NORT-2923BD/2020\|EPI_ISL_489072\|20 | .................... | .................... | .................... | .................... | .................... | .................... |
| E | 4248 | hCoV-19/England/NORT-292DCE/2020\|EPI_ISL_489073\|20 | .................... | .................... | .................... | .................... | .................... | .................... |
| E | 4249 | hCoV-19/England/NORT-2921E0/2020\|EPI_ISL_489074\|20 | .................... | .................... | .................... | .................... | .................... | .................... |
| E | 4250 | hCoV-19/England/NORT-292168/2020\|EPI_ISL_489075\|20 | .................... | .................... | .................... | .................... | .................... | .................... |
| E | 4251 | hCoV-19/England/NORT-292881/2020\|EPI_ISL_489076\|20 | .................... | .................... | .................... | .................... | .................... | .................... |
| E | 4252 | hCoV-19/England/NORT-292256/2020\|EPI_ISL_489077\|20 | .................... | .................... | .................... | .................... | .................... | .................... |
| E | 4253 | hCoV-19/England/NORT-2924E7/2020\|EPI_ISL_489078\|20 | .................... | .................... | .................... | .................... | .................... | .................... |
| E | 4254 | hCoV-19/England/NORT-291F23/2020\|EPI_ISL_489079\|20 | .................... | .................... | .................... | .................... | .................... | .................... |
| E | 4255 | hCoV-19/England/NORT-292C76/2020\|EPI_ISL_489080\|20 | .................... | .................... | .................... | .................... | .................... | .................... |
| E | 4256 | hCoV-19/England/NORT-291E80/2020\|EPI_ISL_489081\|20 | .................... | .................... | .................... | .................... | .................... | .................... |
| E | 4257 | hCoV-19/England/NORT-2922FC/2020\|EPI_ISL_489082\|20 | .................... | .................... | .................... | .................... | .................... | .................... |
| E | 4258 | hCoV-19/England/NORT-29261E/2020\|EPI_ISL_489083\|20 | .................... | .................... | .................... | .................... | .................... | .................... |
| E | 4259 | hCoV-19/England/NORT-292EBC/2020\|EPI_ISL_489084\|20 | .................... | .................... | .................... | .................... | .................... | .................... |
| E | 4260 | hCoV-19/England/NORT-2928AF/2020\|EPI_ISL_489085\|20 | .................... | .................... | .................... | .................... | .................... | .................... |
| E | 4261 | hCoV-19/England/NORT-292274/2020\|EPI_ISL_489086\|20 | .................... | .................... | .................... | .................... | .................... | .................... |
| E | 4262 | hCoV-19/England/NORT-291E53/2020\|EPI_ISL_489087\|20 | .................... | .................... | .................... | .................... | .................... | .................... |
| E | 4263 | hCoV-19/England/NORT-291FD8/2020\|EPI_ISL_489088\|20 | .................... | .................... | .................... | .................... | .................... | .................... |
| E | 4264 | hCoV-19/England/NORT-2929AC/2020\|EPI_ISL_489089\|20 | .................... | .................... | .................... | .................... | .................... | .................... |
| E | 4265 | hCoV-19/England/NORT-292B00/2020\|EPI_ISL_489091\|20 | .................... | .................... | .................... | .................... | .................... | .................... |
| E | 4266 | hCoV-19/England/NORT-29272A/2020\|EPI_ISL_489092\|20 | .................... | .................... | .................... | .................... | .................... | .................... |
| E | 4267 | hCoV-19/England/NORT-292DBF/2020\|EPI_ISL_489093\|20 | .................... | .................... | .................... | .................... | .................... | .................... |
| E | 4268 | hCoV-19/England/NORT-2929F7/2020\|EPI_ISL_489094\|20 | .................... | .................... | .................... | .................... | .................... | .................... |
| E | 4269 | hCoV-19/England/NORT-292669/2020\|EPI_ISL_489097\|20 | .................... | .................... | .................... | .................... | .................... | .................... |
| E | 4270 | hCoV-19/England/NORT-2928BE/2020\|EPI_ISL_489099\|20 | .................... | .................... | .................... | .................... | .................... | .................... |
| E | 4271 | hCoV-19/England/NORT-292380/2020\|EPI_ISL_489100\|20 | .................... | .................... | .................... | .................... | .................... | .................... |
| E | 4272 | hCoV-19/England/NORT-29264B/2020\|EPI_ISL_489101\|20 | .................... | .................... | .................... | .................... | .................... | .................... |
| E | 4273 | hCoV-19/England/NORT-292D37/2020\|EPI_ISL_489102\|20 | .................... | .................... | .................... | .................... | .................... | .................... |
| E | 4274 | hCoV-19/England/NORT-2921FF/2020\|EPI_ISL_489103\|20 | .................... | .................... | .................... | .................... | .................... | .................... |
| E | 4275 | hCoV-19/England/NORT-292EDA/2020\|EPI_ISL_489104\|20 | .................... | .................... | .................... | .................... | .................... | .................... |
| E | 4276 | hCoV-19/England/NORT-292D19/2020\|EPI_ISL_489105\|20 | .................... | .................... | .................... | .................... | .................... | .................... |
| E | 4277 | hCoV-19/Singapore/602/2020\|EPI_ISL_490052\|2020-05- | .................... | .................... | .................... | .................... | .................... | .................... |
| E | 4278 | hCoV-19/Singapore/603/2020\|EPI_ISL_490053\|2020-06- | .................... | .................... | .................... | .................... | .................... | .................... |
| E | 4279 | hCoV-19/Singapore/604/2020\|EPI_ISL_490054\|2020-06- | .................... | .................... | .................... | .................... | .................... | .................... |
| E | 4280 | hCoV-19/Singapore/607/2020\|EPI_ISL_490057\|2020-07- | .................... | .................... | .................... | .................... | .................... | .................... |
| E | 4281 | hCoV-19/Singapore/608/2020\|EPI_ISL_490058\|2020-03- | .................... | .................... | .................... | .................... | .................... | .................... |
| E | 4282 | hCoV-19/Singapore/610/2020\|EPI_ISL_490060\|2020-05- | .................... | .................... | .................... | .................... | N................... | .................... |
| E | 4283 | hCoV-19/Singapore/611/2020\|EPI_ISL_490061\|2020-05- | .................... | .................... | .................... | .................... | .................... | .................... |
| E | 4284 | hCoV-19/Singapore/612/2020\|EPI_ISL_490062\|2020-05- | .................... | .................... | .................... | .................... | .................... | .................... |
| E | 4285 | hCoV-19/Singapore/613/2020\|EPI_ISL_490063\|2020-05- | .................... | .................... | .................... | .................... | .................... | .................... |
| E | 4286 | hCoV-19/Singapore/614/2020\|EPI_ISL_490064\|2020-05- | .................... | .................... | .................... | .................... | .................... | .................... |
| E | 4287 | hCoV-19/Singapore/615/2020\|EPI_ISL_490065\|2020-05- | .................... | .................... | .................... | .................... | .................... | .................... |
| E | 4288 | hCoV-19/Singapore/616/2020\|EPI_ISL_490066\|2020-05- | .................... | .................... | .................... | .................... | .................... | .................... |
| E | 4289 | hCoV-19/Singapore/617/2020\|EPI_ISL_490067\|2020-05- | .................... | .................... | .................... | .................... | .................... | .................... |
| E | 4290 | hCoV-19/Singapore/618/2020\|EPI_ISL_490068\|2020-05- | .................... | .................... | .................... | .................... | .................... | .................... |
| E | 4291 | hCoV-19/Singapore/619/2020\|EPI_ISL_490069\|2020-05- | .................... | .................... | .................... | .................... | .................... | .................... |
| E | 4292 | hCoV-19/Singapore/620/2020\|EPI_ISL_490070\|2020-05- | .................... | .................... | .................... | .................... | .................... | .................... |
| E | 4293 | hCoV-19/Singapore/621/2020\|EPI_ISL_490071\|2020-04- | .................... | .................... | .................... | .................... | .................... | .................... |
| E | 4294 | hCoV-19/Singapore/622/2020\|EPI_ISL_490072\|2020-05- | .................... | .................... | .................... | .................... | .................... | .................... |
| E | 4295 | hCoV-19/Singapore/623/2020\|EPI_ISL_490073\|2020-05- | .................... | .................... | .................... | .................... | .................... | .................... |
| E | 4296 | hCoV-19/Singapore/624/2020\|EPI_ISL_490074\|2020-04- | .................... | .................... | .................... | .................... | .................... | .................... |
| E | 4297 | hCoV-19/Singapore/625/2020\|EPI_ISL_490075\|2020-05- | .................... | .................... | .................... | .................... | .................... | .................... |
| E | 4298 | hCoV-19/Singapore/626/2020\|EPI_ISL_490076\|2020-05- | .................... | .................... | .................... | .................... | .................... | .................... |

| | | | | | | | | | |
|---|---|---|---|---|---|---|---|---|---|
| E | 4299 | hCoV-19/Singapore/627/2020\|EPI_ISL_490077\|2020-05- | ···················· | ···················· | ····················· | ···················· | ···················· | ···················· | |
| E | 4300 | hCoV-19/Singapore/628/2020\|EPI_ISL_490078\|2020-05- | ···················· | ···················· | ····················· | ···················· | ···················· | ···················· | |
| E | 4302 | hCoV-19/Malaysia/IMR_WC55122/2020\|EPI_ISL_490089\|2 | ···················· | ···················· | ····················· | ···················· | ···················· | ···················· | |
| E | 4303 | hCoV-19/Malaysia/IMR_WC80031/2020\|EPI_ISL_490090\|2 | ···················· | ···················· | ····················· | ···················· | ···················· | ···················· | |
| E | 4304 | hCoV-19/Malaysia/IMR_WC80036/2020\|EPI_ISL_490091\|2 | ···················· | ···················· | ····················· | ···················· | ···················· | ···················· | |
| E | 4305 | hCoV-19/Malaysia/IMR_WC80066/2020\|EPI_ISL_490092\|2 | ···················· | ···················· | ····················· | ···················· | ···················· | ···················· | |
| E | 4306 | hCoV-19/Malaysia/IMR_WC80109/2020\|EPI_ISL_490094\|2 | ···················· | ···················· | ····················· | ···················· | ···················· | ···················· | |
| E | 4307 | hCoV-19/Malaysia/IMR_WC80558/2020\|EPI_ISL_490095\|2 | ···················· | ···················· | ····················· | ···················· | ···················· | ···················· | |
| E | 4308 | hCoV-19/Malaysia/IMR_WC81523/2020\|EPI_ISL_490096\|2 | ···················· | ···················· | ····················· | ···················· | ···················· | ···················· | |
| E | 4309 | hCoV-19/Malaysia/IMR_WC81849/2020\|EPI_ISL_490098\|2 | ···················· | ···················· | ····················· | ···················· | ···················· | ···················· | |
| E | 4310 | hCoV-19/Malaysia/IMR_WC99023/2020\|EPI_ISL_490099\|2 | ···················· | ···················· | ····················· | ···················· | ···················· | ···················· | |
| E | 4311 | hCoV-19/USA/WI-UW-705/2020\|EPI_ISL_491383\|2020-06- | ···················· | ···················· | ····················· | ···················· | ···················· | ···················· | |
| E | 4312 | hCoV-19/USA/WI-UW-706/2020\|EPI_ISL_491384\|2020-06- | ···················· | ···················· | ····················· | ···················· | ···················· | ···················· | |
| E | 4313 | hCoV-19/USA/WI-UW-708/2020\|EPI_ISL_491385\|2020-07- | ···················· | ···················· | ····················· | ···················· | ···················· | ···················· | |
| E | 4314 | hCoV-19/USA/WI-UW-709/2020\|EPI_ISL_491386\|2020-06- | ···················· | ···················· | ····················· | ···················· | ···················· | ···················· | |
| E | 4315 | hCoV-19/USA/WI-UW-711/2020\|EPI_ISL_491387\|2020-06- | ···················· | ···················· | ····················· | ···················· | ···················· | ···················· | |
| E | 4316 | hCoV-19/USA/WI-UW-712/2020\|EPI_ISL_491388\|2020-07- | ···················· | ···················· | ····················· | ···················· | ···················· | ···················· | |
| E | 4317 | hCoV-19/USA/WI-UW-713/2020\|EPI_ISL_491389\|2020-06- | ···················· | ···················· | ····················· | ···················· | ···················· | ···················· | |
| E | 4318 | hCoV-19/USA/WI-UW-714/2020\|EPI_ISL_491390\|2020-06- | ···················· | ···················· | ····················· | ···················· | ···················· | ···················· | |
| E | 4319 | hCoV-19/USA/WI-UW-715/2020\|EPI_ISL_491391\|2020-06- | ···················· | ···················· | ····················· | ···················· | ···················· | ···················· | |
| E | 4321 | hCoV-19/USA/WI-UW-719/2020\|EPI_ISL_491394\|2020-06- | ···················· | ···················· | ····················· | ···················· | ···················· | ···················· | |
| E | 4322 | hCoV-19/USA/WI-UW-720/2020\|EPI_ISL_491395\|2020-06- | ···················· | ···················· | ····················· | ···················· | ···················· | ···················· | |
| E | 4323 | hCoV-19/USA/WI-UW-723/2020\|EPI_ISL_491398\|2020-06- | ···················· | ···················· | ····················· | ···················· | ···················· | ···················· | |
| E | 4324 | hCoV-19/USA/WI-UW-724/2020\|EPI_ISL_491399\|2020-07- | ···················· | ···················· | ····················· | ···················· | ···················· | ···················· | |
| E | 4325 | hCoV-19/USA/WI-UW-726/2020\|EPI_ISL_491401\|2020-06- | ···················· | ···················· | ····················· | ···················· | ···················· | ···················· | |
| E | 4326 | hCoV-19/USA/WI-UW-727/2020\|EPI_ISL_491402\|2020-06- | ···················· | ···················· | ····················· | ···················· | ···················· | ···················· | |
| E | 4327 | hCoV-19/USA/WI-UW-728/2020\|EPI_ISL_491403\|2020-07- | ···················· | ···················· | ····················· | ···················· | ···················· | ···················· | |
| E | 4328 | hCoV-19/USA/WI-UW-729/2020\|EPI_ISL_491404\|2020-07- | ···················· | ···················· | ····················· | ···················· | ···················· | ···················· | |
| E | 4329 | hCoV-19/USA/WI-UW-730/2020\|EPI_ISL_491405\|2020-06- | ···················· | ···················· | ····················· | ···················· | ···················· | ···················· | |
| E | 4330 | hCoV-19/USA/WI-UW-731/2020\|EPI_ISL_491406\|2020-07- | ···················· | ···················· | ····················· | ···················· | ···················· | ···················· | |
| E | 4331 | hCoV-19/USA/WI-UW-732/2020\|EPI_ISL_491407\|2020-06- | ···················· | ···················· | ····················· | ···················· | ···················· | ···················· | |
| E | 4332 | hCoV-19/USA/WI-UW-733/2020\|EPI_ISL_491408\|2020-06- | ···················· | ···················· | ····················· | ···················· | ···················· | ···················· | |
| E | 4333 | hCoV-19/USA/WI-UW-734/2020\|EPI_ISL_491409\|2020-07- | ···················· | ···················· | ····················· | ···················· | ···················· | ···················· | |
| E | 4334 | hCoV-19/England/BRIS-130033/2020\|EPI_ISL_492700\|20 | ···················· | ···················· | ····················· | ···················· | ···················· | ···················· | |
| E | 4335 | hCoV-19/England/BRIS-12FE3D/2020\|EPI_ISL_492701\|20 | ···················· | ···················· | ····················· | ···················· | ···················· | ···················· | |
| E | 4336 | hCoV-19/England/BRIS-1306D7/2020\|EPI_ISL_492702\|20 | ···················· | ···················· | ····················· | ···················· | ···················· | ···················· | |
| E | 4337 | hCoV-19/England/BRIS-13022E/2020\|EPI_ISL_492703\|20 | ···················· | ············A······· | ····················· | ···················· | ···················· | ···················· | |
| E | 4338 | hCoV-19/England/BRIS-1301F4/2020\|EPI_ISL_492705\|20 | ···················· | ············A······· | ····················· | ···················· | ···················· | ···················· | |
| E | 4339 | hCoV-19/England/BRIS-12FF76/2020\|EPI_ISL_492706\|20 | ···················· | ···················· | ····················· | ···················· | ···················· | ···················· | |
| E | 4340 | hCoV-19/England/BRIS-12FEF1/2020\|EPI_ISL_492707\|20 | ···················· | ·A·················· | ····················· | ···················· | ···················· | ···················· | |
| E | 4341 | hCoV-19/England/BRIS-1302A6/2020\|EPI_ISL_492708\|20 | ···················· | ············A······· | ····················· | ···················· | ···················· | ···················· | |
| E | 4342 | hCoV-19/England/BRIS-13018B/2020\|EPI_ISL_492709\|20 | ···················· | ···················· | ····················· | ···················· | ···················· | ···················· | |
| E | 4343 | hCoV-19/England/BRIS-130AEA/2020\|EPI_ISL_492730\|20 | ···················· | ···················· | ····················· | ···················· | ···················· | ···················· | |
| E | 4344 | hCoV-19/England/BRIS-130552/2020\|EPI_ISL_492731\|20 | ···················· | ···················· | ····················· | ···················· | ···················· | ···················· | |
| E | 4345 | hCoV-19/England/BRIS-12FE1F/2020\|EPI_ISL_492732\|20 | ···················· | ···················· | ····················· | ···················· | ···················· | ···················· | |
| E | 4346 | hCoV-19/England/BRIS-12FD5E/2020\|EPI_ISL_492733\|20 | ···················· | ···················· | ····················· | ···················· | ···················· | ···················· | |
| E | 4347 | hCoV-19/England/BRIS-1305BC/2020\|EPI_ISL_492734\|20 | ···················· | ···················· | ····················· | ···················· | ···················· | ···················· | |
| E | 4348 | hCoV-19/England/BRIS-130B9C/2020\|EPI_ISL_492735\|20 | ···················· | ···················· | ····················· | ···················· | ···················· | ···················· | |
| E | 4349 | hCoV-19/England/BRIS-130929/2020\|EPI_ISL_492736\|20 | ···················· | ···················· | ····················· | ···················· | ···················· | ···················· | |
| E | 4350 | hCoV-19/England/BRIS-13068C/2020\|EPI_ISL_492737\|20 | ···················· | ···················· | ····················· | ···················· | ···················· | ···················· | |
| E | 4351 | hCoV-19/England/BRIS-12FBEB/2020\|EPI_ISL_492738\|20 | ···················· | ···················· | ····················· | ···················· | ···················· | ···················· | |
| E | 4352 | hCoV-19/England/BRIS-1303FE/2020\|EPI_ISL_492739\|20 | ···················· | ···················· | ····················· | ···················· | ···················· | ···················· | |
| E | 4358 | hCoV-19/Australia/VIC2029/2020\|EPI_ISL_480657\|2020 | ···················· | ···················· | ····················· | ···················· | ···················· | ···················· | |
| E | 4361 | hCoV-19/Australia/VIC2020/2020\|EPI_ISL_480662\|2020 | ···················· | ···················· | ····················· | ···················· | NNN················· | ···················· | 1 |
| E | 4362 | hCoV-19/Australia/VIC2025/2020\|EPI_ISL_480665\|2020 | ···················· | ···················· | ····················· | ···················· | NNN················· | ···················· | 1 |
| E | 4363 | hCoV-19/Australia/VIC2034/2020\|EPI_ISL_480666\|2020 | ···················· | ···················· | ····················· | ···················· | ···················· | ···················· | |
| E | 4366 | hCoV-19/Australia/VIC1944/2020\|EPI_ISL_480669\|2020 | ···················· | ···················· | ····················· | ···················· | ···················· | ···················· | |
| E | 4367 | hCoV-19/Australia/NT47/2020\|EPI_ISL_480694\|2020 | ···················· | ···················· | ····················· | ···················· | ···················· | ···················· | |
| E | 4369 | hCoV-19/Australia/NT49/2020\|EPI_ISL_480696\|2020 | ···················· | ···················· | ····················· | ···················· | ···················· | ···················· | |
| E | 4370 | hCoV-19/Australia/VIC2086/2020\|EPI_ISL_480698\|2020 | ···················· | ···················· | ····················· | ···················· | ···················· | ···················· | |
| E | 4372 | hCoV-19/England/BRIS-12DD50/2020\|EPI_ISL_481984\|20 | ···················· | ···················· | ····················· | ···················· | ···················· | ···················· | |
| E | 4373 | hCoV-19/England/BRIS-12E2E7/2020\|EPI_ISL_481991\|20 | ···················· | ···················· | ····················· | ···················· | ···················· | ···················· | |
| E | 4374 | hCoV-19/England/BRIS-12DD41/2020\|EPI_ISL_481995\|20 | ···················· | ···················· | ····················· | ···················· | ···················· | ···················· | |
| E | 4375 | hCoV-19/England/BRIS-12DDD8/2020\|EPI_ISL_481996\|20 | ···················· | ···················· | ····················· | ···················· | ···················· | ···················· | |
| E | 4376 | hCoV-19/England/BRIS-12DE20/2020\|EPI_ISL_481998\|20 | ···················· | ···················· | ····················· | ···················· | ···················· | ···················· | |
| E | 4377 | hCoV-19/England/BRIS-12DFD2/2020\|EPI_ISL_481999\|20 | ···················· | ···················· | ····················· | ···················· | ···················· | ···················· | |
| E | 4378 | hCoV-19/Russia/CRIE161143/2020\|EPI_ISL_486820\|2020 | ···················· | ···················· | ····················· | ···················· | ···················· | ···················· | |
| E | 4379 | hCoV-19/Russia/CRIE162398/2020\|EPI_ISL_486821\|2020 | ···················· | ···················· | ····················· | ···················· | ···················· | ···················· | |
| E | 4380 | hCoV-19/Russia/CRIE164980/2020\|EPI_ISL_486824\|2020 | ···················· | ···················· | ····················· | ···················· | ···················· | ···················· | |
| E | 4381 | hCoV-19/Russia/CRIE168693/2020\|EPI_ISL_486825\|2020 | ···················· | ···················· | ····················· | ···················· | ···················· | ···················· | |
| E | 4382 | hCoV-19/Russia/CRIE170180/2020\|EPI_ISL_486826\|2020 | ···················· | ···················· | ····················· | ···················· | ···················· | ···················· | |
| E | 4383 | hCoV-19/Russia/CRIE175960/2020\|EPI_ISL_486827\|2020 | ···················· | ···················· | ····················· | ···················· | ···················· | ···················· | |
| E | 4384 | hCoV-19/Russia/CRIE176675/2020\|EPI_ISL_486828\|2020 | ···················· | ···················· | ····················· | ···················· | ···················· | ···················· | |
| E | 4385 | hCoV-19/Russia/CRIE187963/2020\|EPI_ISL_486829\|2020 | ···················· | ···················· | ····················· | ···················· | ···················· | ···················· | |
| E | 4386 | hCoV-19/Scotland/EDB3092/2020\|EPI_ISL_489000\|2020- | ···················· | ···················A | ····················· | ···················· | ···················· | ···················· | 1 |
| E | 4387 | hCoV-19/Scotland/EDB3078/2020\|EPI_ISL_489001\|2020- | ···················· | ···················· | ····················· | ···················· | ···················· | ···················· | |
| E | 4388 | hCoV-19/Scotland/EDB2745/2020\|EPI_ISL_489002\|2020- | ···················· | ···················· | ····················· | ···················· | ···················· | ···················· | |
| E | 4389 | hCoV-19/Scotland/EDB2785/2020\|EPI_ISL_489003\|2020- | ···················· | ···················· | ····················· | ···················· | ···················· | ···················· | |
| E | 4390 | hCoV-19/Scotland/EDB3035/2020\|EPI_ISL_489004\|2020- | ···················· | ···················A | ····················· | ···················· | ···················· | ···················· | 1 |
| E | 4391 | hCoV-19/Scotland/EDB3004/2020\|EPI_ISL_489005\|2020- | ···················· | ···················· | ····················· | ···················· | ···················· | ···················· | |

| | | | | | | | | |
|---|---|---|---|---|---|---|---|---|
| E | 4392 hCoV-19/Scotland/EDB2691/2020\|EPI_ISL_489006\|2020- | ···················· | ···················· | ···················· | ···················· | ···················· | ···················· | |
| E | 4393 hCoV-19/Scotland/EDB3079/2020\|EPI_ISL_489029\|2020- | ···················· | ···················A | ···················· | ···················· | ···················· | ···················· | 1 |
| E | 4394 hCoV-19/Scotland/EDB2913/2020\|EPI_ISL_489030\|2020- | ···················· | ···················· | ···················· | ···················· | ···················· | ···················· | |
| E | 4395 hCoV-19/Scotland/EDB3108/2020\|EPI_ISL_489031\|2020- | ···················· | ···················· | ···················· | ···················· | ···················· | ···················· | |
| E | 4396 hCoV-19/Scotland/EDB3020/2020\|EPI_ISL_489032\|2020- | ···················· | ···················· | ···················· | ···················· | ···················· | ···················· | |
| E | 4397 hCoV-19/Scotland/EDB2812/2020\|EPI_ISL_489033\|2020- | ···················· | ···················A | ···················· | ···················· | ···················· | ···················· | 1 |
| E | 4398 hCoV-19/Scotland/EDB3201/2020\|EPI_ISL_489034\|2020- | ···················· | ···················· | ···················· | ···················· | ···················· | ···················· | |
| E | 4399 hCoV-19/Scotland/EDB3178/2020\|EPI_ISL_489035\|2020- | ···················· | ···················· | ···················· | ···················· | ···················· | ···················· | |
| E | 4400 hCoV-19/Scotland/EDB3256/2020\|EPI_ISL_489036\|2020- | ···················· | ···················· | ···················· | ···················· | ···················· | ···················· | |
| E | 4401 hCoV-19/Scotland/EDB3144/2020\|EPI_ISL_489037\|2020- | ···················· | ···················· | ···················· | ···················· | ···················· | ···················· | |
| E | 4402 hCoV-19/Scotland/EDB3032/2020\|EPI_ISL_489038\|2020- | ···················· | ···················· | ···················· | ···················· | ···················· | ···················· | |
| E | 4403 hCoV-19/Scotland/EDB2810/2020\|EPI_ISL_489039\|2020- | ···················· | ···················· | ···················· | ···················· | ···················· | ···················· | |
| E | 4404 hCoV-19/Scotland/EDB3176/2020\|EPI_ISL_489040\|2020- | ···················· | ···················· | ···················· | ···················· | ···················· | ···················· | |
| E | 4405 hCoV-19/Scotland/EDB2854/2020\|EPI_ISL_489041\|2020- | ···················· | ···················· | ···················· | ···················· | ···················· | ···················· | |
| E | 4406 hCoV-19/Scotland/EDB3206/2020\|EPI_ISL_489042\|2020- | ···················· | ···················· | ···················· | ···················· | ···················· | ···················· | |
| E | 4407 hCoV-19/Scotland/EDB3290/2020\|EPI_ISL_489044\|2020- | ···················· | ···················A | ···················· | ···················· | ···················· | ···················· | 1 |
| E | 4408 hCoV-19/Scotland/EDB3063/2020\|EPI_ISL_489045\|2020- | ···················· | ···················· | ···················· | ···················· | ···················· | ···················· | |
| E | 4409 hCoV-19/Scotland/EDB2989/2020\|EPI_ISL_489046\|2020- | ···················· | ···················· | ···················· | ···················· | ···················· | ···················· | |
| E | 4410 hCoV-19/Scotland/EDB2708/2020\|EPI_ISL_489047\|2020- | ···················· | ···················· | ···················· | ···················· | ···················· | ···················· | |
| E | 4411 hCoV-19/Scotland/EDB3175/2020\|EPI_ISL_489048\|2020- | ···················· | ···················· | ···················· | ···················· | ···················· | ···················· | |
| E | 4412 hCoV-19/Scotland/EDB3305/2020\|EPI_ISL_489049\|2020- | ···················· | ···················· | ···················· | ···················· | ···················· | ···················· | |
| E | 4413 hCoV-19/Scotland/EDB3041/2020\|EPI_ISL_489050\|2020- | ···················· | ···················· | ···················· | ···················· | ···················· | ···················· | |
| E | 4414 hCoV-19/Scotland/EDB3299/2020\|EPI_ISL_489051\|2020- | ···················· | ···················A | ···················· | ···················· | ···················· | ···················· | 1 |
| E | 4415 hCoV-19/Scotland/EDB3040/2020\|EPI_ISL_489052\|2020- | ···················· | ···················· | ···················· | ···················· | ···················· | ···················· | |
| E | 4416 hCoV-19/Scotland/EDB3104/2020\|EPI_ISL_489053\|2020- | ···················· | ···················A | ···················· | ···················· | ···················· | ···················· | 1 |
| E | 4417 hCoV-19/Scotland/EDB2694/2020\|EPI_ISL_489054\|2020- | ···················· | ···················· | ···················· | ···················· | ···················· | ···················· | |
| E | 4418 hCoV-19/Scotland/EDB3225/2020\|EPI_ISL_489055\|2020- | ···················· | ···················· | ···················· | ···················· | ···················· | ···················· | |
| E | 4419 hCoV-19/Scotland/EDB3191/2020\|EPI_ISL_489056\|2020- | ···················· | ···················· | ···················· | ···················· | ···················· | ···················· | |
| E | 4420 hCoV-19/Scotland/EDB2953/2020\|EPI_ISL_489057\|2020- | ···················· | ···················· | ···················· | ···················· | ···················· | ···················· | |
| E | 4421 hCoV-19/Scotland/EDB3166/2020\|EPI_ISL_489058\|2020- | ···················· | ···················A | ···················· | ···················· | ···················· | ···················· | 1 |
| E | 4422 hCoV-19/Scotland/EDB3289/2020\|EPI_ISL_489059\|2020- | ···················· | ···················· | ···················· | ···················· | ···················· | ···················· | |
| E | 4423 hCoV-19/Scotland/EDB2801/2020\|EPI_ISL_489060\|2020- | ···················· | ···················· | ···················· | ···················· | ···················· | ···················· | |
| E | 4424 hCoV-19/Scotland/EDB3028/2020\|EPI_ISL_489061\|2020- | ···················· | ···················A | ···················· | ···················· | ···················· | ···················· | 1 |
| E | 4425 hCoV-19/Scotland/EDB2763/2020\|EPI_ISL_489062\|2020- | ···················· | ···················· | ···················· | ···················· | ···················· | ···················· | |
| E | 4426 hCoV-19/Scotland/EDB3157/2020\|EPI_ISL_489063\|2020- | ···················· | ···················· | ···················· | ···················· | ···················· | ···················· | |
| E | 4427 hCoV-19/England/NORT-291FF6/2020\|EPI_ISL_489065\|20 | ···················· | ···················· | ···················· | ···················· | ···················· | ···················· | |
| E | 4428 hCoV-19/England/NORT-292A6D/2020\|EPI_ISL_489067\|20 | ···················· | ···················· | ···················· | ···················· | ···················· | ···················· | |
| E | 4429 hCoV-19/England/NORT-291DDE/2020\|EPI_ISL_489068\|20 | ···················· | ···················· | ···················· | ···················· | ···················· | ···················· | |
| E | 4430 hCoV-19/Saudi Arabia/523/2020\|EPI_ISL_490000\|2020- | ···················· | ···················· | ···················· | ···················· | ···················· | ···················· | |
| E | 4431 hCoV-19/Saudi Arabia/545/2020\|EPI_ISL_490001\|2020- | ···················· | ···················· | ···················· | ···················· | ···················· | ···A················ | |
| E | 4432 hCoV-19/Saudi Arabia/553/2020\|EPI_ISL_490002\|2020- | ···················· | ···················· | ···················· | ···················· | ···················· | ···················· | |
| E | 4433 hCoV-19/Saudi Arabia/562/2020\|EPI_ISL_490003\|2020- | ···················· | ·A·················· | ···················· | ···················· | ···················· | ···A················ | |
| E | 4434 hCoV-19/Saudi Arabia/656/2020\|EPI_ISL_490010\|2020- | ···················· | ···················· | ···················· | ···················· | ···················· | ···················· | |
| E | 4435 hCoV-19/Saudi Arabia/671/2020\|EPI_ISL_490011\|2020- | ···················· | ···················· | ···················· | ···················· | ···················· | ···················· | |
| E | 4436 hCoV-19/Saudi Arabia/677/2020\|EPI_ISL_490012\|2020- | ···················· | ···················· | ···················· | ···················· | ···················· | ···················· | |
| E | 4437 hCoV-19/Malaysia/IMR_WC10195/2020\|EPI_ISL_490014\|2 | ···················· | ···················· | ···················· | ···················· | ···················· | ···················· | |
| E | 4438 hCoV-19/Malaysia/IMR_WC12286/2020\|EPI_ISL_490016\|2 | ···················· | ···················· | ···················· | ···················· | ···················· | ···················· | |
| E | 4439 hCoV-19/Australia/NSW607/2020\|EPI_ISL_490017\|2020- | ···················· | ···················· | ···················· | ···················· | ···················· | ···················· | |
| E | 4440 hCoV-19/Australia/NSW608/2020\|EPI_ISL_490018\|2020- | ···················· | ···················· | ···················· | ···················· | ···················· | ···················· | |
| E | 4441 hCoV-19/Australia/NSW610/2020\|EPI_ISL_490019\|2020- | ···················· | ···················· | ···················· | ···················· | ···················· | ···················· | |
| E | 4442 hCoV-19/Australia/NSW611/2020\|EPI_ISL_490020\|2020- | ···················· | ···················· | ···················· | ···················· | ···················· | ···················· | |
| E | 4443 hCoV-19/Australia/NSW612/2020\|EPI_ISL_490021\|2020- | ···················· | ···················· | ···················· | ···················· | ···················· | ···················· | |
| E | 4444 hCoV-19/Australia/NSW613/2020\|EPI_ISL_490022\|2020- | ···················· | ···················· | ···················· | ···················· | ···················· | ···················· | |
| E | 4445 hCoV-19/Australia/NSW614/2020\|EPI_ISL_490023\|2020- | ···················· | ···················· | ···················· | ···················· | ···················· | ···················· | |
| E | 4446 hCoV-19/Australia/NSW615/2020\|EPI_ISL_490024\|2020- | ···················· | ·A·················· | ···················· | ···················· | ···················· | ···A················ | |
| E | 4447 hCoV-19/Australia/NSW616/2020\|EPI_ISL_490025\|2020- | ···················· | ···················· | ···················· | ···················· | ···················· | ···················· | |
| E | 4448 hCoV-19/Australia/NSW617/2020\|EPI_ISL_490026\|2020- | ···················· | ···················· | ···················· | ···················· | ···················· | ···················· | |
| E | 4449 hCoV-19/Australia/NSW618/2020\|EPI_ISL_490027\|2020- | ···················· | ···················· | ···················· | ···················· | ···················· | ···················· | |
| E | 4450 hCoV-19/Australia/NSW619/2020\|EPI_ISL_490028\|2020- | ···················· | ···················· | ···················· | ···················· | ···················· | ···················· | |
| E | 4451 hCoV-19/Australia/NSW620/2020\|EPI_ISL_490029\|2020- | ···················· | ···················· | ···················· | ···················· | ···················· | ···················· | |
| E | 4452 hCoV-19/Australia/NSW621/2020\|EPI_ISL_490030\|2020- | ···················· | ···················· | ···················· | ···················· | ···················· | ···················· | |
| E | 4453 hCoV-19/Australia/NSW622/2020\|EPI_ISL_490031\|2020- | ···················· | ···················· | ···················· | ···················· | ···················· | ···················· | |
| E | 4454 hCoV-19/Australia/NSW623/2020\|EPI_ISL_490032\|2020- | ···················· | ···················· | ···················· | ···················· | ···················· | ···················· | |
| E | 4455 hCoV-19/Australia/NSW624/2020\|EPI_ISL_490033\|2020- | ···················· | ···················· | ···················· | ···················· | ···················· | ···················· | |
| E | 4456 hCoV-19/Australia/NSW625/2020\|EPI_ISL_490034\|2020- | ···················· | ···················· | ···················· | ···················· | ···················· | ···················· | |
| E | 4457 hCoV-19/Australia/NSW626/2020\|EPI_ISL_490035\|2020- | ···················· | ···················· | ···················· | ···················· | ···················· | ···················· | |
| E | 4458 hCoV-19/Australia/NSW627/2020\|EPI_ISL_490036\|2020- | ···················· | ···················· | ···················· | ···················· | ···················· | ···················· | |
| E | 4459 hCoV-19/Australia/NSW628/2020\|EPI_ISL_490037\|2020- | ···················· | ···················· | ···················· | ···················· | ···················· | ···················· | |
| E | 4460 hCoV-19/Australia/NSW629/2020\|EPI_ISL_490038\|2020- | ···················· | ···················· | ···················· | ···················· | ···················· | ···················· | |
| E | 4461 hCoV-19/Australia/NSW630/2020\|EPI_ISL_490039\|2020- | ···················· | ···················· | ···················· | ···················· | ···················· | ···················· | |
| E | 4462 hCoV-19/Australia/NSW631/2020\|EPI_ISL_490040\|2020- | ···················· | ···················· | ···················· | ···················· | ···················· | ···················· | |
| E | 4463 hCoV-19/Australia/NSW632/2020\|EPI_ISL_490041\|2020- | ···················· | ···················· | ···················· | ···················· | ···················· | ···················· | |
| E | 4464 hCoV-19/Australia/NSW633/2020\|EPI_ISL_490042\|2020- | ···················· | ·A·················· | ···················· | ···················· | ···················· | ···A················ | |
| E | 4465 hCoV-19/Australia/NSW634/2020\|EPI_ISL_490043\|2020- | ···················· | ···················· | ···················· | ···················· | ···················· | ···················· | |
| E | 4466 hCoV-19/Australia/NSW635/2020\|EPI_ISL_490044\|2020- | ···················· | ···················· | ···················· | ···················· | ···················· | ···················· | |
| E | 4467 hCoV-19/Australia/NSW636/2020\|EPI_ISL_490045\|2020- | ···················· | ···················· | ···················· | ···················· | ···················· | ···················· | |
| E | 4468 hCoV-19/Australia/NSW637/2020\|EPI_ISL_490046\|2020- | ···················· | ···················· | ···················· | ···················· | ···················· | ···················· | |
| E | 4469 hCoV-19/Malaysia/IMR_WC13946/2020\|EPI_ISL_490047\|2 | ···················· | ···················· | ···················· | ···················· | ···················· | ···················· | |
| E | 4470 hCoV-19/Malaysia/IMR_WC14227/2020\|EPI_ISL_490048\|2 | ···················· | ···················· | ···················· | ···················· | ···················· | ···················· | |
| E | 4471 hCoV-19/Singapore/599/2020\|EPI_ISL_490049\|2020-06- | ···················· | ···················· | ···················· | ···················· | ···················· | ···················· | |

| | | | | | | | | |
|---|---|---|---|---|---|---|---|---|
| E | 4472 | hCoV-19/USA/CA-UW-654/2020\|EPI_ISL_491300\|2020-06- | .................... | .................... | .................... | .................... | .................... | .................... |
| E | 4473 | hCoV-19/USA/CA-UW-693/2020\|EPI_ISL_491301\|2020-06- | .................... | .................... | .................... | .................... | .................... | .................... |
| E | 4474 | hCoV-19/USA/CA-UW-710/2020\|EPI_ISL_491302\|2020-06- | .................... | .................... | .................... | .................... | .................... | .................... |
| E | 4475 | hCoV-19/USA/CO-UW-739/2020\|EPI_ISL_491304\|2020-06- | .................... | .................... | .................... | .................... | .................... | .................... |
| E | 4476 | hCoV-19/USA/FL-UW-671/2020\|EPI_ISL_491305\|2020-06- | .................... | .................... | .................... | .................... | .................... | .................... |
| E | 4477 | hCoV-19/USA/FL-UW-741/2020\|EPI_ISL_491306\|2020-06- | .................... | .................... | .................... | .................... | .................... | .................... |
| E | 4478 | hCoV-19/USA/IA-UW-662/2020\|EPI_ISL_491307\|2020-06- | .................... | .................... | .................... | .................... | .................... | .................... |
| E | 4479 | hCoV-19/USA/IL-UW-627/2020\|EPI_ISL_491308\|2020-06- | .................... | .................... | .................... | .................... | .................... | .................... |
| E | 4480 | hCoV-19/USA/IL-UW-639/2020\|EPI_ISL_491309\|2020-06- | .................... | .................... | .................... | .................... | .................... | .................... |
| E | 4481 | hCoV-19/USA/IL-UW-665/2020\|EPI_ISL_491310\|2020-06- | .................... | .................... | .................... | .................... | .................... | .................... |
| E | 4482 | hCoV-19/USA/IL-UW-670/2020\|EPI_ISL_491311\|2020-06- | .................... | .................... | .................... | .................... | .................... | .................... |
| E | 4483 | hCoV-19/USA/IL-UW-686/2020\|EPI_ISL_491312\|2020-06- | .................... | .................... | .................... | .................... | .................... | .................... |
| E | 4484 | hCoV-19/USA/IL-UW-690/2020\|EPI_ISL_491313\|2020-06- | .................... | .................... | .................... | .................... | .................... | .................... |
| E | 4485 | hCoV-19/USA/IL-UW-707/2020\|EPI_ISL_491314\|2020-06- | .................... | .................... | .................... | .................... | .................... | .................... |
| E | 4486 | hCoV-19/USA/MA-UW-633/2020\|EPI_ISL_491316\|2020-06- | .................... | .................... | .................... | .................... | .................... | .................... |
| E | 4487 | hCoV-19/USA/MA-UW-673/2020\|EPI_ISL_491317\|2020-06- | .................... | .................... | .................... | .................... | .................... | .................... |
| E | 4488 | hCoV-19/USA/MN-UW-672/2020\|EPI_ISL_491318\|2020-06- | .................... | .................... | .................... | .................... | .................... | .................... |
| E | 4489 | hCoV-19/USA/NJ-UW-652/2020\|EPI_ISL_491319\|2020-06- | .................... | .................... | .................... | .................... | .................... | .................... |
| E | 4490 | hCoV-19/USA/WI-UW-637/2020\|EPI_ISL_491330\|2020-06- | .................... | .................... | .................... | .................... | .................... | .................... |
| E | 4491 | hCoV-19/USA/WI-UW-638/2020\|EPI_ISL_491331\|2020-06- | .................... | .................... | .................... | .................... | .................... | .................... |
| E | 4492 | hCoV-19/USA/WI-UW-640/2020\|EPI_ISL_491332\|2020-06- | .................... | .................... | .................... | .................... | .................... | .................... |
| E | 4493 | hCoV-19/USA/WI-UW-642/2020\|EPI_ISL_491333\|2020-06- | .................... | .................... | .................... | .................... | .................... | .................... |
| E | 4494 | hCoV-19/USA/WI-UW-643/2020\|EPI_ISL_491334\|2020-06- | .................... | .................... | .................... | .................... | .................... | .................... |
| E | 4495 | hCoV-19/USA/WI-UW-649/2020\|EPI_ISL_491340\|2020-06- | .................... | .................... | .................... | .................... | .................... | .................... |
| E | 4496 | hCoV-19/USA/WI-UW-650/2020\|EPI_ISL_491341\|2020-06- | .................... | .................... | .................... | .................... | .................... | .................... |
| E | 4497 | hCoV-19/USA/WI-UW-653/2020\|EPI_ISL_491342\|2020-06- | .................... | .................... | .................... | .................... | .................... | .................... |
| E | 4498 | hCoV-19/USA/WI-UW-656/2020\|EPI_ISL_491343\|2020-06- | .................... | .................... | .................... | .................... | .................... | .................... |
| E | 4499 | hCoV-19/USA/WI-UW-657/2020\|EPI_ISL_491344\|2020-06- | .................... | .................... | .................... | .................... | .................... | .................... |
| E | 4500 | hCoV-19/USA/WI-UW-658/2020\|EPI_ISL_491345\|2020-06- | .................... | .................... | .................... | .................... | .................... | .................... |
| E | 4501 | hCoV-19/USA/WI-UW-659/2020\|EPI_ISL_491346\|2020-06- | .................... | .................... | .................... | .................... | .................... | .................... |
| E | 4502 | hCoV-19/USA/WI-UW-660/2020\|EPI_ISL_491347\|2020-06- | .................... | .................... | .................... | .................... | .................... | .................... |
| E | 4503 | hCoV-19/USA/WI-UW-661/2020\|EPI_ISL_491348\|2020-06- | .................... | .................... | .................... | .................... | .................... | .................... |
| E | 4504 | hCoV-19/USA/WI-UW-663/2020\|EPI_ISL_491349\|2020-06- | .................... | .................... | .................... | .................... | .................... | .................... |
| E | 4505 | hCoV-19/USA/WI-UW-664/2020\|EPI_ISL_491350\|2020-06- | .................... | .................... | .................... | .................... | .................... | .................... |
| E | 4506 | hCoV-19/USA/WI-UW-666/2020\|EPI_ISL_491351\|2020-06- | .................... | .................... | .................... | .................... | .................... | .................... |
| E | 4507 | hCoV-19/USA/WI-UW-667/2020\|EPI_ISL_491352\|2020-06- | .................... | .................... | .................... | .................... | .................... | .................... |
| E | 4508 | hCoV-19/USA/WI-UW-668/2020\|EPI_ISL_491353\|2020-06- | .................... | .................... | .................... | .................... | .................... | .................... |
| E | 4509 | hCoV-19/USA/WI-UW-669/2020\|EPI_ISL_491354\|2020-06- | .................... | .................... | .................... | .................... | .................... | .................... |
| E | 4510 | hCoV-19/USA/WI-UW-674/2020\|EPI_ISL_491355\|2020-06- | .................... | .................... | .................... | .................... | .................... | .................... |
| E | 4511 | hCoV-19/USA/WI-UW-675/2020\|EPI_ISL_491356\|2020-06- | .................... | .................... | .................... | .................... | .................... | .................... |
| E | 4512 | hCoV-19/USA/WI-UW-676/2020\|EPI_ISL_491357\|2020-06- | .................... | .................... | .................... | .................... | .................... | .................... |
| E | 4513 | hCoV-19/USA/WI-UW-677/2020\|EPI_ISL_491358\|2020-06- | .................... | .................... | .................... | .................... | .................... | .................... |
| E | 4514 | hCoV-19/USA/WI-UW-678/2020\|EPI_ISL_491359\|2020-06- | .................... | .................... | .................... | .................... | .................... | .................... |
| E | 4515 | hCoV-19/USA/WI-UW-679/2020\|EPI_ISL_491360\|2020-06- | .................... | .................... | .................... | .................... | .................... | .................... |
| E | 4516 | hCoV-19/USA/WI-UW-680/2020\|EPI_ISL_491361\|2020-06- | .................... | .................... | .................... | .................... | .................... | .................... |
| E | 4517 | hCoV-19/USA/WI-UW-681/2020\|EPI_ISL_491362\|2020-06- | .................... | .................... | .................... | .................... | .................... | .................... |
| E | 4518 | hCoV-19/USA/WI-UW-682/2020\|EPI_ISL_491363\|2020-06- | .................... | .................... | .................... | .................... | .................... | .................... |
| E | 4519 | hCoV-19/USA/WI-UW-683/2020\|EPI_ISL_491364\|2020-06- | .................... | .................... | .................... | .................... | .................... | .................... |
| E | 4520 | hCoV-19/USA/WI-UW-684/2020\|EPI_ISL_491365\|2020-06- | .................... | .................... | .................... | .................... | .................... | .................... |
| E | 4521 | hCoV-19/USA/WI-UW-685/2020\|EPI_ISL_491366\|2020-06- | .................... | .................... | .................... | .................... | .................... | .................... |
| E | 4522 | hCoV-19/USA/WI-UW-687/2020\|EPI_ISL_491367\|2020-06- | .................... | .................... | .................... | .................... | .................... | .................... |
| E | 4523 | hCoV-19/USA/WI-UW-688/2020\|EPI_ISL_491368\|2020-06- | .................... | .................... | .................... | .................... | .................... | .................... |
| E | 4524 | hCoV-19/USA/WI-UW-689/2020\|EPI_ISL_491369\|2020-06- | .................... | .................... | .................... | .................... | .................... | .................... |
| E | 4525 | hCoV-19/USA/WI-UW-692/2020\|EPI_ISL_491371\|2020-06- | .................... | .................... | .................... | .................... | .................... | .................... |
| E | 4526 | hCoV-19/USA/WI-UW-695/2020\|EPI_ISL_491373\|2020-06- | .................... | .................... | .................... | .................... | .................... | .................... |
| E | 4527 | hCoV-19/USA/WI-UW-696/2020\|EPI_ISL_491374\|2020-07- | .................... | .................... | .................... | .................... | .................... | .................... |
| E | 4528 | hCoV-19/USA/WI-UW-697/2020\|EPI_ISL_491375\|2020-07- | .................... | .................... | .................... | .................... | .................... | .................... |
| E | 4529 | hCoV-19/USA/WI-UW-698/2020\|EPI_ISL_491376\|2020-06- | .................... | .................... | .................... | .................... | .................... | .................... |
| E | 4530 | hCoV-19/USA/WI-UW-699/2020\|EPI_ISL_491377\|2020-06- | .................... | .................... | .................... | .................... | .................... | .................... |
| E | 4531 | hCoV-19/USA/WI-UW-700/2020\|EPI_ISL_491378\|2020-07- | .................... | .................... | .................... | .................... | .................... | .................... |
| E | 4532 | hCoV-19/USA/WI-UW-701/2020\|EPI_ISL_491379\|2020-07- | .................... | .................... | .................... | .................... | .................... | .................... |
| E | 4533 | hCoV-19/USA/WI-UW-702/2020\|EPI_ISL_491380\|2020-06- | .................... | .................... | .................... | .................... | .................... | .................... |
| E | 4534 | hCoV-19/USA/WI-UW-703/2020\|EPI_ISL_491381\|2020-06- | .................... | .................... | .................... | .................... | .................... | .................... |
| E | 4535 | hCoV-19/USA/WI-UW-704/2020\|EPI_ISL_491382\|2020-07- | .................... | .................... | .................... | .................... | .................... | .................... |
| E | 4536 | hCoV-19/England/BRIS-1309CF/2020\|EPI_ISL_492630\|20 | .................... | .................... | .................... | .................... | .................... | .................... |
| E | 4537 | hCoV-19/England/BRIS-13065F/2020\|EPI_ISL_492632\|20 | .................... | .................... | .................... | .................... | .................... | .................... |
| E | 4538 | hCoV-19/England/BRIS-12FBDC/2020\|EPI_ISL_492633\|20 | .................... | .................... | .................... | .................... | .................... | .................... |
| E | 4539 | hCoV-19/England/BRIS-130C5D/2020\|EPI_ISL_492634\|20 | .................... | .................... | .................... | .................... | .................... | .................... |
| E | 4540 | hCoV-19/England/BRIS-12FC42/2020\|EPI_ISL_492635\|20 | .................... | .................... | .................... | .................... | .................... | .................... |
| E | 4541 | hCoV-19/England/BRIS-12FBCD/2020\|EPI_ISL_492636\|20 | .................... | .................... | .................... | .................... | .................... | .................... |
| E | 4542 | hCoV-19/England/BRIS-1306C8/2020\|EPI_ISL_492637\|20 | .................... | .................... | .................... | .................... | .................... | .................... |
| E | 4543 | hCoV-19/England/BRIS-12FC24/2020\|EPI_ISL_492638\|20 | .................... | .................... | .................... | .................... | .................... | .................... |
| E | 4544 | hCoV-19/England/BRIS-12FC8E/2020\|EPI_ISL_492639\|20 | .................... | .................... | .................... | .................... | .................... | .................... |
| E | 4545 | hCoV-19/England/BRIS-12FEC4/2020\|EPI_ISL_492640\|20 | .................... | .................... | .................... | .................... | .................... | .................... |
| E | 4546 | hCoV-19/England/BRIS-1307D4/2020\|EPI_ISL_492641\|20 | .................... | .................... | .................... | .................... | .................... | .................... |
| E | 4547 | hCoV-19/England/BRIS-130D1E/2020\|EPI_ISL_492642\|20 | .................... | .................... | .................... | .................... | .................... | .................... |
| E | 4548 | hCoV-19/England/BRIS-1307F2/2020\|EPI_ISL_492643\|20 | .................... | .................... | .................... | .................... | .................... | .................... |
| E | 4549 | hCoV-19/England/BRIS-13023D/2020\|EPI_ISL_492644\|20 | .................... | .................... | .................... | .................... | .................... | .................... |
| E | 4550 | hCoV-19/England/BRIS-13015E/2020\|EPI_ISL_492645\|20 | .................... | .................... | .................... | .................... | .................... | .................... |
| E | 4551 | hCoV-19/England/BRIS-1302E2/2020\|EPI_ISL_492647\|20 | .................... | .................... | .................... | .................... | .................... | .................... |

| | | | | | | | | |
|---|---|---|---|---|---|---|---|---|
| E | 4552 | hCoV-19/England/BRIS-130455/2020\|EPI_ISL_492648\|20 | ···················· | ···················· | ···········Y········ | ···················· | ···················· | ···················· |
| E | 4553 | hCoV-19/England/BRIS-130051/2020\|EPI_ISL_492649\|20 | ···················· | ·············A······ | ···················· | ···················· | ···················· | ···················· |
| E | 4554 | hCoV-19/England/BRIS-130D4B/2020\|EPI_ISL_492660\|20 | ···················· | ···················· | ···················· | ···················· | ···················· | ···················· |
| E | 4555 | hCoV-19/England/BRIS-130367/2020\|EPI_ISL_492661\|20 | ···················· | ···················· | ···················· | ···················· | ···················· | ···················· |
| E | 4556 | hCoV-19/England/BRIS-130938/2020\|EPI_ISL_492662\|20 | ···················· | ···················· | ···················· | ···················· | ···················· | ···················· |
| E | 4557 | hCoV-19/England/BRIS-1300BB/2020\|EPI_ISL_492663\|20 | ···················· | ···················· | ···················· | ···················· | ···················· | ···················· |
| E | 4558 | hCoV-19/England/BRIS-130AF9/2020\|EPI_ISL_492664\|20 | ···················· | ···················· | ···················· | ···················· | ···················· | ···················· |
| E | 4559 | hCoV-19/England/BRIS-1301E5/2020\|EPI_ISL_492666\|20 | ···················· | ·············A······ | ···················· | ···················· | ···················· | ···················· |
| E | 4560 | hCoV-19/England/BRIS-12FD4F/2020\|EPI_ISL_492670\|20 | ···················· | ···················· | ···················· | ···················· | ···················· | ···················· |
| E | 4561 | hCoV-19/England/BRIS-12FCE8/2020\|EPI_ISL_492671\|20 | ···················· | ···················· | ···················· | ···················· | ···················· | ···················· |
| E | 4562 | hCoV-19/England/BRIS-12FEA6/2020\|EPI_ISL_492673\|20 | ···················· | ···················· | ···················· | ···················· | ···················· | ···················· |
| E | 4563 | hCoV-19/England/BRIS-13069B/2020\|EPI_ISL_492674\|20 | ···················· | ···················· | ···················· | ···················· | ···················· | ···················· |
| E | 4564 | hCoV-19/England/BRIS-130B32/2020\|EPI_ISL_492675\|20 | ···················· | ···················· | ···················· | ···················· | ···················· | ···················· |
| E | 4565 | hCoV-19/England/BRIS-130CD5/2020\|EPI_ISL_492676\|20 | ···················· | ···················· | ···················· | ···················· | ···················· | ···················· |
| E | 4566 | hCoV-19/England/BRIS-12FD7C/2020\|EPI_ISL_492677\|20 | ···················· | ···················· | ···················· | ···················· | ···················· | ···················· |
| E | 4567 | hCoV-19/England/BRIS-12FE6A/2020\|EPI_ISL_492678\|20 | ···················· | ···················· | ···················· | ···················· | ···················· | ···················· |
| E | 4568 | hCoV-19/England/BRIS-1303B2/2020\|EPI_ISL_492679\|20 | ···················· | ···················· | ···················· | ···················· | ···················· | ···················· |
| E | 4569 | hCoV-19/England/BRIS-130622/2020\|EPI_ISL_492680\|20 | ···················· | ···················· | ···················· | ···················· | ···················· | ···················· |
| E | 4570 | hCoV-19/England/BRIS-1300AC/2020\|EPI_ISL_492681\|20 | ···················· | ···················· | ···················· | ···················· | ···················· | ···················· |
| E | 4571 | hCoV-19/England/BRIS-130CF3/2020\|EPI_ISL_492682\|20 | ···················· | ···················· | ···················· | ···················· | ···················· | ···················· |
| E | 4572 | hCoV-19/England/BRIS-130CE4/2020\|EPI_ISL_492683\|20 | ···················· | ···················· | ···················· | ···················· | ···················· | ···················· |
| E | 4573 | hCoV-19/England/BRIS-130516/2020\|EPI_ISL_492684\|20 | ···················· | ···················· | ···················· | ···················· | ···················· | ···················· |
| E | 4574 | hCoV-19/England/BRIS-13040A/2020\|EPI_ISL_492685\|20 | ···················· | ···················· | ···················· | ···················· | ···················· | ···················· |
| E | 4575 | hCoV-19/England/BRIS-13019A/2020\|EPI_ISL_492686\|20 | ···················· | ···················· | ···················· | ···················· | ···················· | ···················· |
| E | 4576 | hCoV-19/England/BRIS-130482/2020\|EPI_ISL_492687\|20 | ···················· | ···················· | ···················· | ···················· | ···················· | ···················· |
| E | 4577 | hCoV-19/England/BRIS-12FC9D/2020\|EPI_ISL_492688\|20 | ···················· | ·A·················· | ···················· | ···················· | ···················· | ···················· |
| E | 4578 | hCoV-19/England/BRIS-12FCCA/2020\|EPI_ISL_492689\|20 | ···················· | ···················· | ···················· | ···················· | ···················· | ···················· |
| E | 4579 | hCoV-19/England/BRIS-1302B5/2020\|EPI_ISL_492690\|20 | ···················· | ····A··············· | ···················· | ···················· | ···················· | ···················· |
| E | 4580 | hCoV-19/England/BRIS-12FE97/2020\|EPI_ISL_492691\|20 | ···················· | ···················· | ···················· | ···················· | ···················· | ···················· |
| E | 4581 | hCoV-19/England/BRIS-1300D9/2020\|EPI_ISL_492692\|20 | ···················· | ···················· | ···················· | ···················· | ···················· | ···················· |
| E | 4582 | hCoV-19/England/BRIS-130525/2020\|EPI_ISL_492693\|20 | ···················· | ···················· | ···················· | ···················· | ···················· | ···················· |
| E | 4583 | hCoV-19/England/BRIS-1309ED/2020\|EPI_ISL_492694\|20 | ···················· | ···················· | ···················· | ···················· | ···················· | ···················· |
| E | 4584 | hCoV-19/England/BRIS-12FE79/2020\|EPI_ISL_492695\|20 | ···················· | ···················· | ···················· | ···················· | ···················· | ···················· |
| E | 4585 | hCoV-19/England/BRIS-13076B/2020\|EPI_ISL_492696\|20 | ···················· | ···················· | ···················· | ···················· | ···················· | ···················· |
| E | 4586 | hCoV-19/England/BRIS-130200/2020\|EPI_ISL_492697\|20 | ···················· | ·············A······ | ···················· | ···················· | ···················· | ···················· |
| E | 4587 | hCoV-19/England/BRIS-13059E/2020\|EPI_ISL_492698\|20 | ···················· | ···················· | ···················· | ···················· | ···················· | ···················· |
| E | 4588 | hCoV-19/England/BRIS-130042/2020\|EPI_ISL_492699\|20 | ···················· | ···················· | ···················· | ···················· | ···················· | ···················· |
| E | 4589 | hCoV-19/Scotland/EDB5621/2020\|EPI_ISL_493960\|2020- | ···················· | ······A············· | ···················· | ···················· | ···················· | ···················· |
| E | 4590 | hCoV-19/Scotland/EDB6640/2020\|EPI_ISL_493963\|2020- | ···················· | ·A·················· | ···················· | ···················· | ···················· | ···A················ |
| E | 4591 | hCoV-19/Scotland/EDB6641/2020\|EPI_ISL_493964\|2020- | ···················· | ···················· | ···················· | ···················· | ···················· | ···················· |
| E | 4592 | hCoV-19/Scotland/EDB6643/2020\|EPI_ISL_493965\|2020- | ···················· | ···················· | ···················· | ···················· | ···················· | ···················· |
| E | 4593 | hCoV-19/Scotland/EDB6646/2020\|EPI_ISL_493968\|2020- | ···················· | ···················· | ···················· | ···················· | ···················· | ···················· |
| E | 4594 | hCoV-19/Scotland/EDB6647/2020\|EPI_ISL_493969\|2020- | ···················· | ···················· | ···················· | ···················· | ···················· | ···················· |
| E | 4595 | hCoV-19/Scotland/EDB6651/2020\|EPI_ISL_493970\|2020- | ···················· | ···················· | ···················· | ···················· | ···················· | ···················· |
| E | 4596 | hCoV-19/Scotland/EDB6652/2020\|EPI_ISL_493971\|2020- | ···················· | ···················· | ···················· | ···················· | ···················· | ···················· |
| E | 4597 | hCoV-19/Scotland/EDB6653/2020\|EPI_ISL_493972\|2020- | ···················· | ···················· | ···················· | ···················· | ···················· | ···················· |
| E | 4598 | hCoV-19/Scotland/EDB6654/2020\|EPI_ISL_493973\|2020- | ···················· | ···················· | ···················· | ···················· | ···················· | ···················· |
| E | 4599 | hCoV-19/Scotland/EDB6718/2020\|EPI_ISL_493974\|2020- | ···················· | ···················· | ···················· | ···················· | ···················· | ···················· |
| E | 4600 | hCoV-19/Wales/PHWC-15C13E/2020\|EPI_ISL_493975\|2020 | ···················· | ···················· | ···················· | ···················· | ···················· | ···················· |
| E | 4602 | hCoV-19/Wales/PHWC-160ED6/2020\|EPI_ISL_493977\|2020 | ···················· | ···················· | ···················· | ···················· | ···················· | ···················· |
| E | 4603 | hCoV-19/Wales/PHWC-1663A4/2020\|EPI_ISL_493990\|2020 | ···················· | ···················· | ···················· | ···················· | ···················· | ···················· |
| E | 4604 | hCoV-19/Wales/PHWC-1663C2/2020\|EPI_ISL_493992\|2020 | ···················· | ···················· | ···················· | ···················· | ···················· | ···················· |
| E | 4605 | hCoV-19/Wales/PHWC-1663D1/2020\|EPI_ISL_493993\|2020 | ···················· | ···················· | ···················· | ···················· | ···················· | ···················· |
| E | 4609 | hCoV-19/Australia/VIC1948/2020\|EPI_ISL_480672\|2020 | ···················· | ···················· | ···················· | ···················· | ···················· | ···················· |
| E | 4611 | hCoV-19/Australia/VIC1950/2020\|EPI_ISL_480674\|2020 | ···················· | ···················· | ···················· | ···················· | ···················· | ···················· |
| E | 4612 | hCoV-19/Australia/VIC1951/2020\|EPI_ISL_480675\|2020 | ···················· | ···················· | ···················· | ···················· | ···················· | ···················· |
| E | 4615 | hCoV-19/Australia/VIC1954/2020\|EPI_ISL_480678\|2020 | ···················· | ···················· | ···················· | ···················· | ···················· | ···················· |
| E | 4617 | hCoV-19/Australia/VIC1956/2020\|EPI_ISL_480680\|2020 | ···················· | ···················· | ···················· | ···················· | ···················· | ···················· |
| E | 4619 | hCoV-19/Australia/VIC1958/2020\|EPI_ISL_480682\|2020 | ···················· | ···················· | ···················· | ···················· | ·NN················· | ···················· |
| E | 4621 | hCoV-19/Australia/VIC2037/2020\|EPI_ISL_480686\|2020 | ···················· | ···················· | ···················· | ···················· | ···················· | ···················· |
| E | 4623 | hCoV-19/England/BRIS-12F468/2020\|EPI_ISL_488300\|20 | ···················· | ···················· | ···················· | ···················· | ···················· | ···················· |
| E | 4624 | hCoV-19/England/BRIS-12E6EB/2020\|EPI_ISL_488301\|20 | ···················· | ···················· | ···················· | ···················· | ···················· | ···················· |
| E | 4625 | hCoV-19/England/BRIS-12E50C/2020\|EPI_ISL_488302\|20 | ···················· | ···················· | ···················· | ···················· | ···················· | ···················· |
| E | 4626 | hCoV-19/England/BRIS-12F1F8/2020\|EPI_ISL_488303\|20 | ···················· | ···················· | ···················· | ···················· | ···················· | ···················· |
| E | 4627 | hCoV-19/England/BRIS-12E548/2020\|EPI_ISL_488304\|20 | ···················· | ···················· | ···················· | ···················· | ···················· | ···················· |
| E | 4628 | hCoV-19/England/BRIS-12F398/2020\|EPI_ISL_488305\|20 | ···················· | ···················· | ···················· | ···················· | ···················· | ···················· |
| E | 4629 | hCoV-19/England/BRIS-12F7F6/2020\|EPI_ISL_488306\|20 | ···················· | ···················· | ···················· | ···················· | ···················· | ···················· |
| E | 4630 | hCoV-19/England/BRIS-12E645/2020\|EPI_ISL_488307\|20 | ···················· | ·············A······ | ···················· | ···················· | ···················· | ···················· |
| E | 4631 | hCoV-19/England/BRIS-12EA3A/2020\|EPI_ISL_488308\|20 | ···················· | ···················· | ···················· | ···················· | ···················· | ···················· |
| E | 4632 | hCoV-19/England/BRIS-12F4E0/2020\|EPI_ISL_488309\|20 | ···················· | ···················· | ···················· | A··················· | ···················· | ···················· |
| E | 4633 | hCoV-19/England/BRIS-12F6EA/2020\|EPI_ISL_488310\|20 | ···················· | ···················· | ···················· | ···················· | ···················· | ···················· |
| E | 4634 | hCoV-19/England/BRIS-12E4D2/2020\|EPI_ISL_488311\|20 | ···················· | ···················· | ···················· | ···················· | ···················· | ···················· |
| E | 4635 | hCoV-19/England/BRIS-12F978/2020\|EPI_ISL_488312\|20 | ···················· | ···················· | ···················· | ···················· | ···················· | ···················· |
| E | 4636 | hCoV-19/England/BRIS-12EE3E/2020\|EPI_ISL_488314\|20 | ···················· | ···················· | ···················· | ···················· | ···················· | ···················· |
| E | 4637 | hCoV-19/England/BRIS-12FA2A/2020\|EPI_ISL_488315\|20 | ···················· | ···················· | ···················· | ···················· | ···················· | ···················· |
| E | 4638 | hCoV-19/England/BRIS-12F028/2020\|EPI_ISL_488316\|20 | ···················· | ···················· | ···················· | ···················· | ···················· | ···················· |
| E | 4639 | hCoV-19/England/BRIS-12EAC1/2020\|EPI_ISL_488317\|20 | ···················· | ···················· | ···················· | ···················· | ···················· | ···················· |
| E | 4640 | hCoV-19/England/BRIS-12EB55/2020\|EPI_ISL_488318\|20 | ···················· | ····A··············· | ···················· | ···················· | ···················· | ···················· |
| E | 4641 | hCoV-19/England/BRIS-12EC16/2020\|EPI_ISL_488319\|20 | ···················· | ···················· | ···················· | ···················· | ···················· | ···················· |
| E | 4642 | hCoV-19/England/BRIS-12F161/2020\|EPI_ISL_488321\|20 | ···················· | ···················· | ···················· | ···················· | ···················· | ···················· |

| | | | | | | | | | |
|---|---|---|---|---|---|---|---|---|---|
| E | 4643 | hCoV-19/England/BRIS-12EEB6/2020\|EPI_ISL_488322\|20 | ···················· | ·············A······ | ···················· | ···················· | ···················· | ···················· | |
| E | 4644 | hCoV-19/England/BRIS-12ED9B/2020\|EPI_ISL_488323\|20 | ···················· | ···················· | ···················· | ···················· | ···················· | ···················· | |
| E | 4645 | hCoV-19/England/BRIS-12F608/2020\|EPI_ISL_488324\|20 | ···················· | ···················· | ···················· | ···················· | ···················· | ···················· | |
| E | 4646 | hCoV-19/England/BRIS-12F671/2020\|EPI_ISL_488337\|20 | ···················· | ···················· | ···················· | ···················· | ···················· | ···················· | |
| E | 4647 | hCoV-19/England/BRIS-12EE89/2020\|EPI_ISL_488338\|20 | ···················· | ···················· | ···················· | ···················· | ···················· | ···················· | |
| E | 4648 | hCoV-19/England/BRIS-12EA67/2020\|EPI_ISL_488339\|20 | ···················· | ···················· | ···················· | ···················· | ···················· | ···················· | |
| E | 4649 | hCoV-19/England/BRIS-12FA57/2020\|EPI_ISL_488341\|20 | ···················· | ···················· | ···················· | ···················· | ···················· | ···················· | |
| E | 4650 | hCoV-19/England/BRIS-12EAD0/2020\|EPI_ISL_488343\|20 | ···················· | ···················· | ···················· | ···················· | ···················· | ···················· | |
| E | 4651 | hCoV-19/England/BRIS-12F43B/2020\|EPI_ISL_488345\|20 | ···················· | ·············A······ | ···················· | ···················· | ···················· | ···················· | |
| E | 4652 | hCoV-19/England/BRIS-12EFE0/2020\|EPI_ISL_488346\|20 | ···················· | ···················· | ···················· | ···················· | ···················· | ···················· | |
| E | 4653 | hCoV-19/England/BRIS-12FB18/2020\|EPI_ISL_488347\|20 | ···················· | ···················· | ···················· | ···················· | ···················· | ···················· | |
| E | 4654 | hCoV-19/England/BRIS-12E91F/2020\|EPI_ISL_488348\|20 | ···················· | ···················· | ···················· | ···················· | ···················· | ···················· | |
| E | 4655 | hCoV-19/England/BRIS-12F750/2020\|EPI_ISL_488349\|20 | ···················· | ···················· | ···················· | ···················· | ···················· | ···················· | |
| E | 4656 | hCoV-19/England/BRIS-12F547/2020\|EPI_ISL_488351\|20 | ···················· | ···················· | ···················· | ···················· | ···················· | ···················· | |
| E | 4657 | hCoV-19/England/BRIS-12ECDA/2020\|EPI_ISL_488352\|20 | ···················· | ···················· | ···················· | ···················· | ···················· | ···················· | |
| E | 4658 | hCoV-19/England/BRIS-12EE01/2020\|EPI_ISL_488353\|20 | ···················· | ···················· | ···················· | ···················· | ···················· | ···················· | |
| E | 4659 | hCoV-19/England/BRIS-12E654/2020\|EPI_ISL_488354\|20 | ···················· | ···················· | ···················· | ···················· | ···················· | ···················· | |
| E | 4660 | hCoV-19/England/BRIS-12F69F/2020\|EPI_ISL_488355\|20 | ···················· | ···················· | ···················· | ···················· | ···················· | ···················· | |
| E | 4661 | hCoV-19/England/BRIS-12F802/2020\|EPI_ISL_488356\|20 | ···················· | ···················· | ···················· | ···················· | ···················· | ···················· | |
| E | 4662 | hCoV-19/England/BRIS-12EA49/2020\|EPI_ISL_488357\|20 | ···················· | ···················· | ···················· | ···················· | ···················· | ···················· | |
| E | 4663 | hCoV-19/England/BRIS-12F222/2020\|EPI_ISL_488358\|20 | ···················· | ···M················ | ···················· | ···················· | ···················· | ···················· | |
| E | 4664 | hCoV-19/England/BRIS-12F231/2020\|EPI_ISL_488359\|20 | ···················· | ···················· | ···················· | ···················· | ···················· | ···················· | |
| E | 4665 | hCoV-19/England/BRIS-12EB64/2020\|EPI_ISL_488360\|20 | ···················· | ···················· | ···················· | ···················· | ···················· | ···················· | |
| E | 4666 | hCoV-19/England/BRIS-12F2F5/2020\|EPI_ISL_488361\|20 | ···················· | ···················· | ···················· | ···················· | ···················· | ···················· | |
| E | 4667 | hCoV-19/England/BRIS-12F8E4/2020\|EPI_ISL_488362\|20 | ···················· | ···················· | ···················· | ···················· | ···················· | ···················· | |
| E | 4668 | hCoV-19/England/BRIS-12F143/2020\|EPI_ISL_488363\|20 | ···················· | ···················· | ···················· | ···················· | ···················· | ···················· | |
| E | 4669 | hCoV-19/England/BRIS-12F6AE/2020\|EPI_ISL_488364\|20 | ···················· | ···················· | ···················· | ···················· | ···················· | ···················· | |
| E | 4670 | hCoV-19/England/BRIS-12F3C5/2020\|EPI_ISL_488365\|20 | ···················· | ···················· | ···················· | ···················· | ···················· | ···················· | |
| E | 4671 | hCoV-19/England/BRIS-12F714/2020\|EPI_ISL_488367\|20 | ···················· | ···················· | ···················· | ···················· | ···················· | ···················· | |
| E | 4672 | hCoV-19/England/BRIS-12E93D/2020\|EPI_ISL_488369\|20 | ···················· | ···················· | ···················· | ···················· | ···················· | ···················· | |
| E | 4673 | hCoV-19/England/BRIS-12F583/2020\|EPI_ISL_488371\|20 | ···················· | ···················· | ···················· | ···················· | ···················· | ···················· | |
| E | 4674 | hCoV-19/England/BRIS-12F26E/2020\|EPI_ISL_488372\|20 | ···················· | ···················· | ···················· | A··················· | ···················· | ···················· | |
| E | 4675 | hCoV-19/England/BRIS-12F25F/2020\|EPI_ISL_488373\|20 | ···················· | ···················· | ···················· | ···················· | ···················· | ···················· | |
| E | 4676 | hCoV-19/England/BRIS-12F1CB/2020\|EPI_ISL_488374\|20 | ···················· | ···················· | ···················· | ···················· | ···················· | ···················· | |
| E | 4677 | hCoV-19/England/BRIS-12E900/2020\|EPI_ISL_488376\|20 | ···················· | ···················· | ···················· | ···················· | ···················· | ···················· | |
| E | 4678 | hCoV-19/England/BRIS-12E803/2020\|EPI_ISL_488377\|20 | ···················· | ···················· | ···················· | ···················· | ···················· | ···················· | |
| E | 4679 | hCoV-19/England/BRIS-12EA2B/2020\|EPI_ISL_488378\|20 | ···················· | ···················· | ···················· | ···················· | ···················· | ···················· | |
| E | 4680 | hCoV-19/England/BRIS-12EBCE/2020\|EPI_ISL_488379\|20 | ···················· | ···················· | ···················· | ···················· | ···················· | ···················· | |
| E | 4681 | hCoV-19/Scotland/EDB3101/2020\|EPI_ISL_489007\|2020- | ···················· | ···················A | ···················· | ···················· | ···················· | ···················· | 1 |
| E | 4682 | hCoV-19/Scotland/EDB3000/2020\|EPI_ISL_489008\|2020- | ···················· | ···················A | ···················· | ···················· | ···················· | ···················· | 1 |
| E | 4683 | hCoV-19/Scotland/EDB3169/2020\|EPI_ISL_489009\|2020- | ···················· | ···················· | ···················· | ···················· | ···················· | ···················· | |
| E | 4684 | hCoV-19/Scotland/EDB2906/2020\|EPI_ISL_489010\|2020- | ···················· | ···················A | ···················· | ···················· | ···················· | ···················· | 1 |
| E | 4685 | hCoV-19/Scotland/EDB3268/2020\|EPI_ISL_489011\|2020- | ···················· | ···················A | ···················· | ···················· | ···················· | ···················· | 1 |
| E | 4686 | hCoV-19/Scotland/EDB3194/2020\|EPI_ISL_489012\|2020- | ···················· | ···················· | ···················· | ···················· | ···················· | ···················· | |
| E | 4687 | hCoV-19/Scotland/EDB2954/2020\|EPI_ISL_489013\|2020- | ···················· | ···················· | ···················· | ···················· | ···················· | ···················· | |
| E | 4688 | hCoV-19/Scotland/EDB3132/2020\|EPI_ISL_489014\|2020- | ···················· | ···················A | ···················· | ···················· | ···················· | ···················· | 1 |
| E | 4689 | hCoV-19/Scotland/EDB2795/2020\|EPI_ISL_489015\|2020- | ···················· | ···················A | ···················· | ···················· | ···················· | ···················· | 1 |
| E | 4690 | hCoV-19/Scotland/EDB2834/2020\|EPI_ISL_489016\|2020- | ···················· | ···················· | ···················· | ···················· | ···················· | ···················· | |
| E | 4691 | hCoV-19/Scotland/EDB3309/2020\|EPI_ISL_489017\|2020- | ···················· | ···················· | ···················· | ···················· | ···················· | ···················· | |
| E | 4692 | hCoV-19/Scotland/EDB2720/2020\|EPI_ISL_489018\|2020- | ···················· | ···················· | ···················· | ···················· | ···················· | ···················· | |
| E | 4693 | hCoV-19/Scotland/EDB3164/2020\|EPI_ISL_489019\|2020- | ···················· | ···················· | ···················· | ···················· | ···················· | ···················· | |
| E | 4694 | hCoV-19/Scotland/EDB2725/2020\|EPI_ISL_489020\|2020- | ···················· | ···················· | ···················· | ···················· | ···················· | ···················· | |
| E | 4695 | hCoV-19/Scotland/EDB3072/2020\|EPI_ISL_489021\|2020- | ···················· | ···················· | ···················· | ···················· | ···················· | ···················· | |
| E | 4696 | hCoV-19/Scotland/EDB3056/2020\|EPI_ISL_489022\|2020- | ···················· | ···················A | ···················· | ···················· | ···················· | ···················· | 1 |
| E | 4697 | hCoV-19/Scotland/EDB2981/2020\|EPI_ISL_489023\|2020- | ···················· | ···················· | ···················· | ···················· | ···················· | ···················· | |
| E | 4698 | hCoV-19/Scotland/EDB3237/2020\|EPI_ISL_489024\|2020- | ···················· | ···················· | ···················· | ···················· | ···················· | ···················· | |
| E | 4699 | hCoV-19/Scotland/EDB2839/2020\|EPI_ISL_489025\|2020- | ···················· | ···················· | ···················· | ···················· | ···················· | ···················· | |
| E | 4700 | hCoV-19/Scotland/EDB2804/2020\|EPI_ISL_489026\|2020- | ···················· | ···················· | ···················· | ···················· | ···················· | ···················· | |
| E | 4701 | hCoV-19/Scotland/EDB2893/2020\|EPI_ISL_489027\|2020- | ···················· | ···················· | ···················· | ···················· | ···················· | ···················· | |
| E | 4702 | hCoV-19/Scotland/EDB2844/2020\|EPI_ISL_489028\|2020- | ···················· | ···················· | ···················· | ···················· | ···················· | ···················· | |
| E | 4703 | hCoV-19/Scotland/GCVR-16F930/2020\|EPI_ISL_489623\|2 | ···················· | ···················· | ···················· | ···················· | ···················· | ···················· | |
| E | 4704 | hCoV-19/Scotland/GCVR-17077E/2020\|EPI_ISL_489625\|2 | ···················· | ···················· | ···················· | ···················· | ···················· | ···················· | |
| E | 4705 | hCoV-19/Scotland/GCVR-16FA2E/2020\|EPI_ISL_489626\|2 | ···················· | ···················· | ···················· | ···················· | ···················· | ···················· | |
| E | 4706 | hCoV-19/Scotland/GCVR-170F0D/2020\|EPI_ISL_489627\|2 | ···················· | ···················· | ···················· | ···················· | ···················· | ···················· | |
| E | 4707 | hCoV-19/Scotland/GCVR-171704/2020\|EPI_ISL_489628\|2 | ···················· | ···················· | ···················· | ···················· | ···················· | ···················· | |
| E | 4708 | hCoV-19/Scotland/GCVR-16FC82/2020\|EPI_ISL_489631\|2 | ···················· | ···················· | ···················· | ···················· | ···················· | ···················· | |
| E | 4709 | hCoV-19/Scotland/GCVR-16FE04/2020\|EPI_ISL_489632\|2 | ···················· | ···················· | ···················· | ···················· | ···················· | ···················· | |
| E | 4710 | hCoV-19/Scotland/GCVR-170750/2020\|EPI_ISL_489633\|2 | ···················· | ···················· | ···················· | ···················· | ···················· | ···················· | |
| E | 4711 | hCoV-19/Scotland/GCVR-16FC0A/2020\|EPI_ISL_489635\|2 | ···················· | ···················A | ···················· | ···················· | ···················· | ···················· | 1 |
| E | 4712 | hCoV-19/Scotland/GCVR-1706EA/2020\|EPI_ISL_489637\|2 | ···················· | ···················· | ···················· | ···················· | ···················· | ···················· | |
| E | 4713 | hCoV-19/Scotland/GCVR-170732/2020\|EPI_ISL_489638\|2 | ···················· | ···················· | ···················· | ···················· | ···················· | ···················· | |
| E | 4714 | hCoV-19/Scotland/GCVR-16FBA3/2020\|EPI_ISL_489639\|2 | ···················· | ···················· | ···················· | ···················· | ···················· | ···················· | |
| E | 4715 | hCoV-19/Scotland/GCVR-17191D/2020\|EPI_ISL_489640\|2 | ···················· | ···················A | ···················· | ···················· | ···················· | ···················· | 1 |
| E | 4716 | hCoV-19/Scotland/GCVR-1707C9/2020\|EPI_ISL_489641\|2 | ···················· | ···················· | ···················· | ···················· | ···················· | ···················· | |
| E | 4717 | hCoV-19/Scotland/GCVR-16FAA6/2020\|EPI_ISL_489642\|2 | ···················· | ·A·················· | ···················· | ···················· | ···················· | ···················· | |
| E | 4718 | hCoV-19/Scotland/GCVR-16FB58/2020\|EPI_ISL_489643\|2 | ···················· | ·A·················· | ···················· | ···················· | ···················· | ···················· | |
| E | 4719 | hCoV-19/Scotland/GCVR-16F912/2020\|EPI_ISL_489647\|2 | ···················· | ···················· | ···················· | ···················· | ···················· | ···················· | |
| E | 4720 | hCoV-19/Scotland/GCVR-16FA5B/2020\|EPI_ISL_489648\|2 | ···················· | ···················· | ···················· | ···················· | ···················· | ···················· | |
| E | 4721 | hCoV-19/Scotland/GCVR-16FB76/2020\|EPI_ISL_489649\|2 | ···················· | ···················A | ···················· | ···················· | ···················· | ···················· | 1 |
| E | 4722 | hCoV-19/Scotland/GCVR-16FDE9/2020\|EPI_ISL_489650\|2 | ···················· | ···················A | ···················· | ···················· | ···················· | ···················· | 1 |

| | | | | | | | | |
|---|---|---|---|---|---|---|---|---|
| E | 4723 hCoV-19/Scotland/GCVR-1707BA/2020\|EPI_ISL_489651\|2 | ···················· | ···················· | ···················· | ···················· | ···················· | ···················· | |
| E | 4724 hCoV-19/Scotland/GCVR-16FE22/2020\|EPI_ISL_489654\|2 | ···················· | ···················· | ···················· | ···················· | ···················· | ···················· | |
| E | 4725 hCoV-19/Scotland/GCVR-1718D4/2020\|EPI_ISL_489661\|2 | ···················· | ···················· | ···················· | ···················· | ···················· | ···················· | |
| E | 4726 hCoV-19/Scotland/GCVR-170714/2020\|EPI_ISL_489663\|2 | ···················· | ···················A | ···················· | ···················· | ···················· | ···················· | 1 |
| E | 4727 hCoV-19/Scotland/GCVR-170ED3/2020\|EPI_ISL_489664\|2 | ···················· | ···················· | ···················· | ···················· | ···················· | ···················· | |
| E | 4728 hCoV-19/Scotland/GCVR-171713/2020\|EPI_ISL_489666\|2 | ···················· | ·A·················· | ···················· | ···················· | ···················· | ···················· | |
| E | 4729 hCoV-19/Scotland/GCVR-16FC73/2020\|EPI_ISL_489667\|2 | ···················· | ···················· | ···················· | ···················· | ···················· | ···················· | |
| E | 4730 hCoV-19/Scotland/GCVR-1707F6/2020\|EPI_ISL_489668\|2 | ···················· | ···················· | ···················· | ···················· | ···················· | ···················· | |
| E | 4731 hCoV-19/Scotland/GCVR-16FD07/2020\|EPI_ISL_489669\|2 | ···················· | ···················A | ···················· | ···················· | ···················· | ···················· | 1 |
| E | 4732 hCoV-19/Scotland/GCVR-16FDCB/2020\|EPI_ISL_489670\|2 | ···················· | ·A·················· | ···················· | ···················· | ···················· | ···················· | |
| E | 4733 hCoV-19/Scotland/GCVR-170802/2020\|EPI_ISL_489672\|2 | ···················· | ·A·················· | ···················· | ···················· | ···················· | ···················· | |
| E | 4734 hCoV-19/Scotland/GCVR-17143A/2020\|EPI_ISL_489674\|2 | ···················· | ···················A | ···················· | ···················· | ···················· | ···················· | 1 |
| E | 4735 hCoV-19/Scotland/GCVR-170EE2/2020\|EPI_ISL_489676\|2 | ···················· | ···················· | ···················· | ···················· | ···················· | ···················· | |
| E | 4736 hCoV-19/Scotland/GCVR-1716DA/2020\|EPI_ISL_489678\|2 | ···················· | ·A·················· | ···················· | ···················· | ···················· | ···················· | |
| E | 4737 hCoV-19/Scotland/GCVR-170E88/2020\|EPI_ISL_489679\|2 | ···················· | ···················· | ···················· | ···················· | ···················· | ···················· | |
| E | 4738 hCoV-19/Scotland/GCVR-17079C/2020\|EPI_ISL_489680\|2 | ···················· | ···················· | ···················· | ···················· | ···················· | ···················· | |
| E | 4739 hCoV-19/Scotland/GCVR-16FCDD/2020\|EPI_ISL_489682\|2 | ···················· | ···················· | ···················· | ···················· | ···················· | ···················· | |
| E | 4740 hCoV-19/Scotland/GCVR-16FE7D/2020\|EPI_ISL_489683\|2 | ···················· | ···················· | ···················· | ···················· | ···················· | ···················· | |
| E | 4741 hCoV-19/Scotland/GCVR-170F3A/2020\|EPI_ISL_489684\|2 | ···················· | ···················· | ···················· | ···················· | ···················· | ···················· | |
| E | 4742 hCoV-19/Scotland/GCVR-1718B6/2020\|EPI_ISL_489685\|2 | ···················· | ···················· | ···················· | ···················· | ···················· | ···················· | |
| E | 4743 hCoV-19/Scotland/GCVR-1714EF/2020\|EPI_ISL_489689\|2 | ···················· | ···················· | ···················· | ···················· | ···················· | ···················· | |
| E | 4744 hCoV-19/Scotland/GCVR-16FCEC/2020\|EPI_ISL_489690\|2 | ···················· | ···················· | ···················· | ···················· | ···················· | ···················· | |
| E | 4745 hCoV-19/Scotland/GCVR-16F9E5/2020\|EPI_ISL_489692\|2 | ···················· | ···················· | ···················· | ···················· | ···················· | ···················· | |
| E | 4746 hCoV-19/Scotland/GCVR-16FF01/2020\|EPI_ISL_489693\|2 | ···················· | ···················· | ···················· | ···················· | ···················· | ···················· | |
| E | 4747 hCoV-19/Scotland/GCVR-16FB94/2020\|EPI_ISL_489694\|2 | ···················· | ···················· | ···················· | ···················· | ···················· | ···················· | |
| E | 4748 hCoV-19/Scotland/GCVR-171546/2020\|EPI_ISL_489697\|2 | ···················· | ···················· | ···················· | ···················· | ···················· | ···················· | |
| E | 4749 hCoV-19/Scotland/GCVR-1717C8/2020\|EPI_ISL_489698\|2 | ···················· | ···················· | ···················· | ···················· | ···················· | ···················· | |
| E | 4750 hCoV-19/Scotland/GCVR-17177D/2020\|EPI_ISL_489699\|2 | ···················· | ···················· | ···················· | ···················· | ···················· | ···················· | |
| E | 4751 hCoV-19/Saudi Arabia/578/2020\|EPI_ISL_490004\|2020- | ···················· | ·A·················· | ···················· | ···················· | ···················· | ··A················· | |
| E | 4752 hCoV-19/Saudi Arabia/583/2020\|EPI_ISL_490005\|2020- | ···················· | ···················· | ···················· | ···················· | ···················· | ···················· | |
| E | 4753 hCoV-19/Saudi Arabia/596/2020\|EPI_ISL_490006\|2020- | ···················· | ···················· | ···················· | ···················· | ···················· | ··A················· | |
| E | 4754 hCoV-19/Saudi Arabia/610/2020\|EPI_ISL_490007\|2020- | ···················· | ·A·················· | ···················· | ···················· | ···················· | ···················· | |
| E | 4755 hCoV-19/Saudi Arabia/627/2020\|EPI_ISL_490008\|2020- | ···················· | ···················· | ···················· | ···················· | ···················· | ···················· | |
| E | 4756 hCoV-19/Saudi Arabia/640/2020\|EPI_ISL_490009\|2020- | ···················· | ···················· | ···················· | ···················· | ···················· | ···················· | |
| E | 4757 hCoV-19/England/SHEF-C8435/2020\|EPI_ISL_490611\|202 | ···················· | ···················· | ···················· | ···················· | ···················· | ···················· | |
| E | 4758 hCoV-19/England/SHEF-C871E/2020\|EPI_ISL_490612\|202 | ···················· | ···················· | ···················· | ···················· | ···················· | ···················· | |
| E | 4759 hCoV-19/England/SHEF-C8839/2020\|EPI_ISL_490613\|202 | ···················· | ···················· | ···················· | ···················· | ···················· | ···················· | |
| E | 4760 hCoV-19/England/SHEF-C8F16/2020\|EPI_ISL_490614\|202 | ···················· | ···················· | ···················· | ···················· | ···················· | ···················· | |
| E | 4761 hCoV-19/England/SHEF-C9328/2020\|EPI_ISL_490615\|202 | ···················· | ···················· | ···················· | ···················· | ···················· | ···················· | |
| E | 4762 hCoV-19/England/SHEF-CA257/2020\|EPI_ISL_490617\|202 | ···················· | ···················· | ···················· | ···················· | ···················· | ···················· | |
| E | 4763 hCoV-19/England/SHEF-CBDCE/2020\|EPI_ISL_490621\|202 | ···················· | ···················· | ···················· | ···················· | ···················· | ···················· | |
| E | 4764 hCoV-19/England/SHEF-CBE43/2020\|EPI_ISL_490622\|202 | ···················· | ···················· | ···················· | ···················· | ···················· | ···················· | |
| E | 4765 hCoV-19/England/SHEF-CC158/2020\|EPI_ISL_490623\|202 | ···················· | ···················· | ···················· | ···················· | ···················· | ···················· | |
| E | 4766 hCoV-19/England/SHEF-CCBB4/2020\|EPI_ISL_490624\|202 | ···················· | ···················· | ···················· | ···················· | ···················· | ···················· | |
| E | 4767 hCoV-19/England/SHEF-CD421/2020\|EPI_ISL_490626\|202 | ···················· | ···················· | ···················· | ···················· | ···················· | ···················· | |
| E | 4768 hCoV-19/England/SHEF-CDEC9/2020\|EPI_ISL_490627\|202 | ···················· | ···················· | ···················· | ···················· | ···················· | ···················· | |
| E | 4769 hCoV-19/England/SHEF-CDF6C/2020\|EPI_ISL_490628\|202 | ···················· | ···················· | ···················· | ···················· | ···················· | ···················· | |
| E | 4770 hCoV-19/England/SHEF-CE280/2020\|EPI_ISL_490629\|202 | ···················· | ···················· | ···················· | ···················· | ···················· | ···················· | |
| E | 4771 hCoV-19/England/SHEF-CE2F9/2020\|EPI_ISL_490630\|202 | ···················· | ···················· | ···················· | ···················· | ···················· | ···················· | |
| E | 4772 hCoV-19/England/SHEF-CFC54/2020\|EPI_ISL_490633\|202 | ···················· | ···················· | ···················· | ···················· | ···················· | ···················· | |
| E | 4773 hCoV-19/England/SHEF-D0018/2020\|EPI_ISL_490635\|202 | ···················· | ···················· | ···················· | ···················· | ···················· | ···················· | |
| E | 4774 hCoV-19/England/SHEF-D0582/2020\|EPI_ISL_490637\|202 | ···················· | ···················· | ···················· | ···················· | ···················· | ···················· | |
| E | 4775 hCoV-19/England/SHEF-D0D11/2020\|EPI_ISL_490638\|202 | ···················· | ···················· | ···················· | ···················· | ···················· | ···················· | |
| E | 4776 hCoV-19/England/SHEF-D177C/2020\|EPI_ISL_490642\|202 | ···················· | ···················· | ···················· | ···················· | ···················· | ···················· | |
| E | 4777 hCoV-19/Scotland/CVR1992/2020\|EPI_ISL_490646\|2020- | ···················· | ·A·················· | ···················· | ···················· | ···················· | ···················· | |
| E | 4780 hCoV-19/Scotland/CVR2041/2020\|EPI_ISL_490654\|2020- | ···················· | ···················· | ···················· | ···················· | ···················· | ···················· | |
| E | 4781 hCoV-19/Scotland/CVR2058/2020\|EPI_ISL_490657\|2020- | ···················· | ···················A | ···················· | ···················· | ···················· | ···················· | 1 |
| E | 4784 hCoV-19/Scotland/CVR2096/2020\|EPI_ISL_490662\|2020- | ···················· | ···················· | ···················· | ···················· | ···················· | ···················· | |
| E | 4785 hCoV-19/Scotland/CVR2101/2020\|EPI_ISL_490664\|2020- | ···················· | ···················A | ···················· | ···················· | ···················· | ···················· | 1 |
| E | 4786 hCoV-19/Scotland/CVR2114/2020\|EPI_ISL_490665\|2020- | ···················· | ···················· | ···················· | ···················· | ···················· | ···················· | |
| E | 4789 hCoV-19/Scotland/CVR2141/2020\|EPI_ISL_490670\|2020- | ···················· | ···················A | ···················· | ···················· | ···················· | ···················· | 1 |
| E | 4790 hCoV-19/Scotland/CVR2232/2020\|EPI_ISL_490675\|2020- | ···················· | ···················A | ···················· | ···················· | ···················· | ···················· | 1 |
| E | 4792 hCoV-19/Scotland/CVR2360/2020\|EPI_ISL_490685\|2020- | ···················· | ···················· | ···················· | ···················· | ···················· | ···················· | |
| E | 4793 hCoV-19/Scotland/CVR2362/2020\|EPI_ISL_490686\|2020- | ···················· | ···················· | ···················· | ···················· | ···················· | ···················· | |
| E | 4795 hCoV-19/USA/NY-UW-641/2020\|EPI_ISL_491320\|2020-06- | ···················· | ···················· | ···················· | ···················· | ···················· | ···················· | |
| E | 4796 hCoV-19/USA/NY-UW-651/2020\|EPI_ISL_491321\|2020-06- | ···················· | ···················· | ···················· | ···················· | ···················· | ···················· | |
| E | 4797 hCoV-19/USA/WI-UW-626/2020\|EPI_ISL_491322\|2020-06- | ···················· | ···················· | ···················· | ···················· | ···················· | ···················· | |
| E | 4798 hCoV-19/USA/WI-UW-628/2020\|EPI_ISL_491323\|2020-06- | ···················· | ···················· | ···················· | ···················· | ···················· | ···················· | |
| E | 4799 hCoV-19/USA/WI-UW-630/2020\|EPI_ISL_491324\|2020-06- | ···················· | ···················· | ···················· | ···················· | ···················· | ···················· | |
| E | 4800 hCoV-19/USA/WI-UW-631/2020\|EPI_ISL_491325\|2020-06- | ···················· | ···················· | ···················· | ···················· | ···················· | ···················· | |
| E | 4801 hCoV-19/USA/WI-UW-632/2020\|EPI_ISL_491326\|2020-06- | ···················· | ···················· | ···················· | ···················· | ···················· | ···················· | |
| E | 4802 hCoV-19/USA/WI-UW-634/2020\|EPI_ISL_491327\|2020-06- | ···················· | ···················· | ···················· | ···················· | ···················· | ···················· | |
| E | 4803 hCoV-19/USA/WI-UW-635/2020\|EPI_ISL_491328\|2020-06- | ···················· | ···················· | ···················· | ···················· | ···················· | ···················· | |
| E | 4804 hCoV-19/USA/WI-UW-644/2020\|EPI_ISL_491335\|2020-06- | ···················· | ···················· | ···················· | ···················· | ···················· | ···················· | |
| E | 4805 hCoV-19/USA/WI-UW-645/2020\|EPI_ISL_491336\|2020-06- | ···················· | ···················· | ···················· | ···················· | ···················· | ···················· | |
| E | 4806 hCoV-19/USA/WI-UW-646/2020\|EPI_ISL_491337\|2020-06- | ···················· | ···················· | ···················· | ···················· | ···················· | ···················· | |
| E | 4807 hCoV-19/USA/WI-UW-647/2020\|EPI_ISL_491338\|2020-06- | ···················· | ···················· | ···················· | ···················· | ···················· | ···················· | |
| E | 4808 hCoV-19/USA/WI-UW-648/2020\|EPI_ISL_491339\|2020-06- | ···················· | ···················· | ···················· | ···················· | ···················· | ···················· | |
| E | 4809 hCoV-19/Ecuador/USFQ-127/2020\|EPI_ISL_491940\|2020- | ···················· | ···················· | ···················· | ···················· | ···················· | ···················· | |
| E | 4810 hCoV-19/Ecuador/USFQ-133/2020\|EPI_ISL_491941\|2020- | ···················· | ···················· | ···················· | ···················· | ···················· | ···················· | |

| | | | | | | | | |
|---|---|---|---|---|---|---|---|---|
| E | 4811 hCoV-19/USA/VA_NIDDL_3082/2020\|EPI_ISL_491942\|2020 | .................... | .................... | .................... | .................... | .................... | .................... | |
| E | 4812 hCoV-19/USA/VA_NIDDL_3216/2020\|EPI_ISL_491943\|2020 | .................... | .................... | .................... | .................... | .................... | .................... | |
| E | 4813 hCoV-19/Ecuador/23890/2020\|EPI_ISL_491944\|2020-04- | .................... | .................... | .................... | .................... | .................... | .................... | |
| E | 4814 hCoV-19/Ecuador/23055/2020\|EPI_ISL_491945\|2020-04- | .................... | .................... | .................... | .................... | .................... | .................... | |
| E | 4815 hCoV-19/Ecuador/52872/2020\|EPI_ISL_491946\|2020-05- | .................... | .................... | .................... | .................... | .................... | .................... | |
| E | 4816 hCoV-19/Ecuador/36192/2020\|EPI_ISL_491947\|2020-05- | .................... | .................... | .................... | .................... | .................... | .................... | |
| E | 4817 hCoV-19/Ecuador/51908/2020\|EPI_ISL_491949\|2020-05- | .................... | .................... | .................... | .................... | .................... | .................... | |
| E | 4818 hCoV-19/Ecuador/36230/2020\|EPI_ISL_491950\|2020-05- | .................... | .................... | .................... | .................... | .................... | .................... | |
| E | 4819 hCoV-19/Ecuador/52438/2020\|EPI_ISL_491951\|2020-06- | .................... | .................... | .................... | .................... | .................... | .................... | |
| E | 4820 hCoV-19/Ecuador/52255/2020\|EPI_ISL_491952\|2020-05- | .................... | .................... | .................... | .................... | .................... | .................... | |
| E | 4821 hCoV-19/Ecuador/36226/2020\|EPI_ISL_491953\|2020-05- | .................... | .................... | .................... | .................... | .................... | .................... | |
| E | 4822 hCoV-19/Ecuador/52389/2020\|EPI_ISL_491954\|2020-05- | .................... | .................... | .................... | .................... | .................... | .................... | |
| E | 4823 hCoV-19/Oman/205035037/2020\|EPI_ISL_491968\|2020-05 | .................... | .................... | .................... | .................... | .................... | .................... | |
| E | 4824 hCoV-19/Oman/205035503/2020\|EPI_ISL_491969\|2020-05 | .................... | .................... | .................... | .................... | .................... | .................... | |
| E | 4825 hCoV-19/Oman/205035515/2020\|EPI_ISL_491970\|2020-05 | .................... | .................... | .................... | .................... | .................... | .................... | |
| E | 4826 hCoV-19/Oman/205036082/2020\|EPI_ISL_491973\|2020-05 | .................... | .................... | .................... | .................... | .................... | .................... | |
| E | 4827 hCoV-19/Oman/205036136/2020\|EPI_ISL_491974\|2020-05 | .................... | .................... | .................... | .................... | .................... | .................... | |
| E | 4828 hCoV-19/Oman/205036179/2020\|EPI_ISL_491976\|2020-05 | .................... | .................... | .................... | .................... | .................... | .................... | |
| E | 4829 hCoV-19/Oman/205036454/2020\|EPI_ISL_491978\|2020-05 | .................... | .................... | .................... | .................... | .................... | .................... | |
| E | 4830 hCoV-19/Oman/205036457/2020\|EPI_ISL_491979\|2020-05 | .................... | .................... | .................... | .................... | .................... | .................... | |
| E | 4831 hCoV-19/Oman/205036471/2020\|EPI_ISL_491980\|2020-05 | .................... | .................... | .................... | .................... | .................... | .................... | |
| E | 4832 hCoV-19/Oman/205036690/2020\|EPI_ISL_491981\|2020-05 | .................... | .................... | .................... | .................... | .................... | .................... | |
| E | 4833 hCoV-19/Oman/205036702/2020\|EPI_ISL_491982\|2020-05 | .................... | .................... | .................... | .................... | .................... | .................... | |
| E | 4834 hCoV-19/Oman/205036713/2020\|EPI_ISL_491983\|2020-05 | .................... | .................... | .................... | .................... | .................... | .................... | |
| E | 4835 hCoV-19/Oman/205036714/2020\|EPI_ISL_491984\|2020-05 | .................... | .................... | .................... | .................... | .................... | .................... | |
| E | 4836 hCoV-19/Oman/205037001/2020\|EPI_ISL_491985\|2020-05 | .................... | .................... | .................... | .................... | .................... | .................... | |
| E | 4837 hCoV-19/Oman/205037397/2020\|EPI_ISL_491986\|2020-05 | .................... | .................... | .................... | .................... | .................... | .................... | |
| E | 4838 hCoV-19/Oman/205037858/2020\|EPI_ISL_491988\|2020-05 | .................... | .................... | .................... | .................... | .................... | .................... | |
| E | 4839 hCoV-19/Oman/205038017/2020\|EPI_ISL_491989\|2020-05 | .................... | .................... | .................... | .................... | .................... | .................... | |
| E | 4840 hCoV-19/Oman/205038033/2020\|EPI_ISL_491990\|2020-05 | .................... | .................... | .................... | .................... | .................... | .................... | |
| E | 4841 hCoV-19/Oman/205038035/2020\|EPI_ISL_491991\|2020-05 | .................... | .................... | .................... | .................... | .................... | .................... | |
| E | 4842 hCoV-19/Oman/205038041/2020\|EPI_ISL_491992\|2020-05 | .................... | .................... | .................... | .................... | .................... | .................... | |
| E | 4843 hCoV-19/Oman/205038170/2020\|EPI_ISL_491994\|2020-05 | .................... | .................... | .................... | .................... | .................... | .................... | |
| E | 4844 hCoV-19/Oman/205038321/2020\|EPI_ISL_491995\|2020-05 | .................... | .................... | .................... | .................... | .................... | .................... | |
| E | 4845 hCoV-19/Oman/205038337/2020\|EPI_ISL_491996\|2020-05 | .................... | .................... | .................... | .................... | .................... | .................... | |
| E | 4846 hCoV-19/Oman/205038687/2020\|EPI_ISL_491997\|2020-05 | .................... | .A.................. | .................... | .................... | .................... | .................... | |
| E | 4847 hCoV-19/Oman/205038852/2020\|EPI_ISL_491998\|2020-05 | .................... | .................... | .................... | .................... | .................... | .................... | |
| E | 4848 hCoV-19/Oman/205038885/2020\|EPI_ISL_491999\|2020-05 | .................... | .................... | .................... | .................... | .................... | .................... | |
| E | 4849 hCoV-19/England/BRIS-130385/2020\|EPI_ISL_492651\|20 | .................... | .................... | .................... | .................... | .................... | .................... | |
| E | 4850 hCoV-19/England/BRIS-12FC7F/2020\|EPI_ISL_492652\|20 | .................... | .................... | .................... | .................... | .................... | .................... | |
| E | 4851 hCoV-19/England/BRIS-130789/2020\|EPI_ISL_492653\|20 | .................... | .................... | .................... | .................... | .................... | .................... | |
| E | 4852 hCoV-19/England/BRIS-13084A/2020\|EPI_ISL_492654\|20 | .................... | .................... | .................... | .................... | .................... | .................... | |
| E | 4853 hCoV-19/England/BRIS-1308E0/2020\|EPI_ISL_492655\|20 | .................... | .................... | .................... | .................... | .................... | .................... | |
| E | 4854 hCoV-19/England/BRIS-130868/2020\|EPI_ISL_492656\|20 | .................... | .................... | .................... | .................... | .................... | .................... | |
| E | 4855 hCoV-19/England/BRIS-1302D3/2020\|EPI_ISL_492657\|20 | .................... | .................... | .................... | .................... | .................... | .................... | |
| E | 4856 hCoV-19/England/BRIS-130561/2020\|EPI_ISL_492658\|20 | .................... | .................... | .................... | .................... | .................... | .................... | |
| E | 4857 hCoV-19/England/BRIS-130BD8/2020\|EPI_ISL_492659\|20 | .................... | .................... | .................... | .................... | .................... | .................... | |
| E | 4858 hCoV-19/England/BRIS-130983/2020\|EPI_ISL_492665\|20 | .................... | .................... | .................... | .................... | .................... | .................... | |
| E | 4859 hCoV-19/England/BRIS-1305DA/2020\|EPI_ISL_492667\|20 | .................... | .................... | .................... | .................... | .................... | .................... | |
| E | 4860 hCoV-19/England/BRIS-13021F/2020\|EPI_ISL_492669\|20 | .................... | .............A...... | .................... | .................... | .................... | .................... | |
| E | 4861 hCoV-19/Wales/PHWC-166359/2020\|EPI_ISL_493985\|2020 | .................... | .................... | .................... | .................... | .................... | .................... | |
| E | 4862 hCoV-19/Wales/PHWC-166377/2020\|EPI_ISL_493987\|2020 | .................... | .................... | .................... | .................... | NNN................. | .................... | 1 |
| E | 4864 hCoV-19/Wales/PHWC-166456/2020\|EPI_ISL_493999\|2020 | .................... | .................... | .................... | .................... | .................... | .................... | |
| E | 4865 hCoV-19/England/HECH-2599D91/2020\|EPI_ISL_482111\|2 | .................... | .................... | .................... | .................... | .................... | .................... | |
| E | 4867 hCoV-19/England/HECH-2599DDD/2020\|EPI_ISL_482113\|2 | .................... | .................... | .................... | .................... | .................... | .................... | |
| E | 4868 hCoV-19/England/HECH-2599EAD/2020\|EPI_ISL_482114\|2 | .................... | .................... | .................... | .................... | .................... | .................... | |
| E | 4869 hCoV-19/Northern Ireland/NIRE-101253/2020\|EPI_ISL_ | .................... | .................... | .................... | .................... | .................... | .................... | |
| E | 4870 hCoV-19/England/HECH-259A0C4/2020\|EPI_ISL_482116\|2 | .................... | .................... | .................... | .................... | .................... | .................... | |
| E | 4871 hCoV-19/England/HECH-2599D28/2020\|EPI_ISL_482117\|2 | .................... | .................... | .................... | .................... | .................... | .................... | |
| E | 4872 hCoV-19/England/HECH-2599FF5/2020\|EPI_ISL_482118\|2 | .................... | .................... | .................... | .................... | .................... | .................... | |
| E | 4873 hCoV-19/England/TBSD-248867D/2020\|EPI_ISL_482120\|2 | .................... | .................... | .................... | .................... | .................... | .................... | |
| E | 4874 hCoV-19/England/TBSD-2488A26/2020\|EPI_ISL_482128\|2 | .................... | .................... | .................... | .................... | .................... | .................... | |
| E | 4875 hCoV-19/England/TBSD-2488A08/2020\|EPI_ISL_482130\|2 | .................... | .................... | .................... | .................... | .................... | .................... | |
| E | 4876 hCoV-19/Northern Ireland/NIRE-1042F6/2020\|EPI_ISL_ | .................... | .................... | .................... | .................... | .................... | .................... | |
| E | 4877 hCoV-19/Northern Ireland/NIRE-103F0F/2020\|EPI_ISL_ | .................... | .................... | .................... | .................... | .................... | .................... | |
| E | 4878 hCoV-19/England/LOND-125F9F3/2020\|EPI_ISL_482138\|2 | .................... | .................... | .................... | .................... | .................... | .................... | |
| E | 4879 hCoV-19/Northern Ireland/NIRE-1016FD/2020\|EPI_ISL_ | .................... | .................... | .................... | .................... | .................... | .................... | |
| E | 4880 hCoV-19/Northern Ireland/NIRE-10110B/2020\|EPI_ISL_ | .................... | .................... | .................... | .................... | .................... | .................... | |
| E | 4881 hCoV-19/Northern Ireland/NIRE-100B68/2020\|EPI_ISL_ | .................... | .................... | .................... | .................... | .................... | .................... | |
| E | 4882 hCoV-19/England/LOND-D62A3/2020\|EPI_ISL_482160\|202 | .................... | .................... | .................... | .................... | .................... | .................... | |
| E | 4883 hCoV-19/England/LOND-D61F1/2020\|EPI_ISL_482161\|202 | .................... | .................... | .................... | .................... | .................... | .................... | |
| E | 4884 hCoV-19/England/LOND-D6100/2020\|EPI_ISL_482163\|202 | .................... | .................... | .................... | .................... | .................... | .................... | |
| E | 4885 hCoV-19/England/LOND-D6865/2020\|EPI_ISL_482165\|202 | .................... | .................... | .................... | .................... | .................... | .................... | |
| E | 4886 hCoV-19/England/LOND-D6768/2020\|EPI_ISL_482166\|202 | .................... | .................... | .................... | .................... | .................... | .................... | |
| E | 4887 hCoV-19/England/LOND-D612E/2020\|EPI_ISL_482168\|202 | .................... | .................... | .................... | .................... | .................... | .................... | |
| E | 4888 hCoV-19/England/LOND-D611F/2020\|EPI_ISL_482170\|202 | .................... | .................... | .................... | .................... | .................... | .................... | |
| E | 4889 hCoV-19/England/LOND-D6373/2020\|EPI_ISL_482171\|202 | .................... | .................... | .................... | .................... | .................... | .................... | |
| E | 4890 hCoV-19/England/LOND-D670E/2020\|EPI_ISL_482175\|202 | .................... | .................... | .................... | .................... | .................... | .................... | |
| E | 4891 hCoV-19/England/LOND-D6786/2020\|EPI_ISL_482176\|202 | .................... | .................... | .................... | .................... | .................... | .................... | |
| E | 4892 hCoV-19/England/LOND-D616A/2020\|EPI_ISL_482178\|202 | .................... | .................... | .................... | .................... | .................... | .................... | |

| | | | | | | | | |
|---|---|---|---|---|---|---|---|---|
| E | 4893 hCoV-19/England/LOND-D6179/2020\|EPI_ISL_482182\|202 | ···················· | ···················· | ···················· | ···················· | ···················· | ···················· | |
| E | 4894 hCoV-19/England/LOND-D6197/2020\|EPI_ISL_482183\|202 | ···················· | ···················· | ···················· | ···················· | ···················· | ···················· | |
| E | 4895 hCoV-19/England/LOND-D6355/2020\|EPI_ISL_482184\|202 | ···················· | ···················· | ···················· | ···················· | ···················· | ···················· | |
| E | 4896 hCoV-19/England/LOND-D67B3/2020\|EPI_ISL_482186\|202 | ···················· | ···················· | ···················· | ···················· | ···················· | ···················· | |
| E | 4897 hCoV-19/England/LOND-D66D4/2020\|EPI_ISL_482187\|202 | ···················· | ···················· | ···················· | ···················· | ···················· | ···················· | |
| E | 4898 hCoV-19/England/LOND-D62B2/2020\|EPI_ISL_482188\|202 | ···················· | ···················· | ···················· | ···················· | ···················· | ···················· | |
| E | 4899 hCoV-19/England/LOND-D6276/2020\|EPI_ISL_482191\|202 | ···················· | ···················· | ···················· | ···················· | ···················· | ···················· | |
| E | 4900 hCoV-19/England/LOND-D6382/2020\|EPI_ISL_482192\|202 | ···················· | ······A············· | ···················· | ···················· | ···················· | ···················· | |
| E | 4901 hCoV-19/England/LOND-D67E0/2020\|EPI_ISL_482196\|202 | ···················· | ···················· | ···················· | ···················· | ···················· | ···················· | |
| E | 4902 hCoV-19/England/LOND-D68ED/2020\|EPI_ISL_482197\|202 | ···················· | ···················· | ···················· | ···················· | ···················· | ···················· | |
| E | 4903 hCoV-19/USA/CA-ALSR-0982-SAN/2020\|EPI_ISL_483441\|2 | ···················· | ···················· | ···················· | ···················· | ···················· | ···················· | |
| E | 4904 hCoV-19/USA/CA-ALSR-0987-SAN/2020\|EPI_ISL_483444\|2 | ···················· | ···················· | ···················· | ···················· | ···················· | ···················· | |
| E | 4905 hCoV-19/USA/CA-ALSR-0989-SAN/2020\|EPI_ISL_483445\|2 | ···················· | ···················· | ···················· | ···················· | ···················· | ···················· | |
| E | 4906 hCoV-19/USA/CA-ALSR-0990-SAN/2020\|EPI_ISL_483446\|2 | ···················· | ···················· | ···················· | ···················· | ···················· | ···················· | |
| E | 4907 hCoV-19/USA/CA-ALSR-0991-SAN/2020\|EPI_ISL_483447\|2 | ···················· | ···················· | ···················· | ···················· | ···················· | ···················· | |
| E | 4908 hCoV-19/USA/CA-ALSR-0993-SAN/2020\|EPI_ISL_483449\|2 | ···················· | ·A·················· | ···················· | ···················· | ···················· | ···················· | |
| E | 4909 hCoV-19/USA/CA-ALSR-0996-IPL/2020\|EPI_ISL_483450\|2 | ···················· | ···················· | ···················· | ···················· | ···················· | ···················· | |
| E | 4910 hCoV-19/USA/CA-ALSR-0997-IPL/2020\|EPI_ISL_483451\|2 | ···················· | ·A·················· | ···················· | ···················· | ···················· | ···················· | |
| E | 4911 hCoV-19/USA/CA-ALSR-0998-IPL/2020\|EPI_ISL_483452\|2 | ···················· | ·A·················· | ···················· | ···················· | ···················· | ···················· | |
| E | 4912 hCoV-19/USA/CA-ALSR-0999-SAN/2020\|EPI_ISL_483453\|2 | ···················· | ·A·················· | ···················· | ···················· | ···················· | ···················· | |
| E | 4913 hCoV-19/USA/CA-ALSR-1000-SAN/2020\|EPI_ISL_483454\|2 | ···················· | ·A·················· | ···················· | ···················· | ···················· | ···················· | |
| E | 4914 hCoV-19/USA/CA-ALSR-1001-IPL/2020\|EPI_ISL_483455\|2 | ···················· | ···················· | ···················· | ···················· | ···················· | ···················· | |
| E | 4915 hCoV-19/USA/CA-ALSR-1002-SAN/2020\|EPI_ISL_483456\|2 | ···················· | ···················· | ···················· | ···················· | ···················· | ···················· | |
| E | 4916 hCoV-19/USA/CA-ALSR-1003-SAN/2020\|EPI_ISL_483457\|2 | ···················· | ·A·················· | ···················· | ···················· | ···················· | ···················· | |
| E | 4917 hCoV-19/USA/CA-ALSR-1004-SAN/2020\|EPI_ISL_483458\|2 | ···················· | ·A·················· | ···················· | ···················· | ···················· | ···················· | |
| E | 4918 hCoV-19/USA/CA-ALSR-1007-SAN/2020\|EPI_ISL_483459\|2 | ···················· | ·A·················· | ···················· | ···················· | ···················· | ···················· | |
| E | 4919 hCoV-19/USA/CA-ALSR-1012-SAN/2020\|EPI_ISL_483460\|2 | ···················· | ···················· | ···················· | ···················· | ···················· | ···················· | |
| E | 4920 hCoV-19/USA/CA-ALSR-1013-SAN/2020\|EPI_ISL_483461\|2 | ···················· | ···················· | ···················· | ···················· | ···················· | ···················· | |
| E | 4921 hCoV-19/USA/CA-ALSR-1014-SAN/2020\|EPI_ISL_483462\|2 | ···················· | ·A·················· | ···················· | ···················· | ···················· | ···················· | |
| E | 4922 hCoV-19/USA/CA-ALSR-1015-IPL/2020\|EPI_ISL_483463\|2 | ···················· | ···················· | ···················· | ···················· | ···················· | ···················· | |
| E | 4923 hCoV-19/USA/CA-ALSR-1016-IPL/2020\|EPI_ISL_483464\|2 | ···················· | ·A·················· | ···················· | ···················· | ···················· | ···················· | |
| E | 4924 hCoV-19/USA/CA-ALSR-1018-SAN/2020\|EPI_ISL_483465\|2 | ···················· | ···················· | ···················· | ···················· | ···················· | ···················· | |
| E | 4925 hCoV-19/USA/CA-ALSR-1019-IPL/2020\|EPI_ISL_483466\|2 | ···················· | ·A·················· | ···················· | ···················· | ···················· | ···················· | |
| E | 4926 hCoV-19/USA/CA-ALSR-1021-IPL/2020\|EPI_ISL_483467\|2 | ···················· | ·A·················· | ···················· | ···················· | ···················· | ···················· | |
| E | 4927 hCoV-19/USA/CA-ALSR-1022-SAN/2020\|EPI_ISL_483468\|2 | ···················· | ·A·················· | ···················· | ···················· | ···················· | ···················· | |
| E | 4928 hCoV-19/USA/CA-ALSR-1023-SAN/2020\|EPI_ISL_483469\|2 | ···················· | ·A·················· | ···················· | ···················· | ···················· | ···················· | |
| E | 4929 hCoV-19/USA/CA-ALSR-1025-IPL/2020\|EPI_ISL_483470\|2 | ···················· | ···················· | ···················· | ···················· | ···················· | ···················· | |
| E | 4930 hCoV-19/USA/CA-ALSR-1027-SAN/2020\|EPI_ISL_483471\|2 | ···················· | ·A·················· | ···················· | ···················· | ···················· | ···················· | |
| E | 4931 hCoV-19/USA/CA-ALSR-1028-IPL/2020\|EPI_ISL_483472\|2 | ···················· | ·A·················· | ···················· | ···················· | ···················· | ···················· | |
| E | 4932 hCoV-19/USA/CA-ALSR-1029-SAN/2020\|EPI_ISL_483473\|2 | ···················· | ···················· | ···················· | ···················· | ···················· | ···················· | |
| E | 4933 hCoV-19/USA/CA-ALSR-1053-SAN/2020\|EPI_ISL_483485\|2 | ···················· | ·A·················· | ···················· | ···················· | ···················· | ···················· | |
| E | 4934 hCoV-19/USA/CA-ALSR-1055-SAN/2020\|EPI_ISL_483486\|2 | ···················· | ·A·················· | ···················· | ···················· | ···················· | ···················· | |
| E | 4935 hCoV-19/USA/CA-ALSR-1056-SAN/2020\|EPI_ISL_483487\|2 | ···················· | ·A·················· | ···················· | ···················· | ···················· | ···················· | |
| E | 4936 hCoV-19/USA/CA-ALSR-1057-SAN/2020\|EPI_ISL_483488\|2 | ···················· | ···················· | ···················· | ···················· | ···················· | ···················· | |
| E | 4937 hCoV-19/USA/CA-ALSR-1058-SAN/2020\|EPI_ISL_483489\|2 | ···················· | ·A·················· | ···················· | ···················· | ···················· | ···················· | |
| E | 4938 hCoV-19/USA/CA-ALSR-1059-SAN/2020\|EPI_ISL_483490\|2 | ···················· | ···················· | ···················· | ···················· | ···················· | ···················· | |
| E | 4939 hCoV-19/USA/CA-ALSR-1060-SAN/2020\|EPI_ISL_483491\|2 | ···················· | ·A·················· | ···················· | ···················· | ···················· | ···················· | |
| E | 4940 hCoV-19/USA/CA-ALSR-1061-SAN/2020\|EPI_ISL_483492\|2 | ···················· | ·A·················· | ···················· | ···················· | ···················· | ···················· | |
| E | 4941 hCoV-19/USA/CA-ALSR-1062-SAN/2020\|EPI_ISL_483493\|2 | ···················· | ·A·················· | ···················· | ···················· | ···················· | ···················· | |
| E | 4942 hCoV-19/USA/CA-ALSR-1063-SAN/2020\|EPI_ISL_483494\|2 | ···················· | ···················· | ···················· | ···················· | ···················· | ···················· | |
| E | 4943 hCoV-19/USA/CA-ALSR-1065-SAN/2020\|EPI_ISL_483495\|2 | ···················· | ·A·················· | ···················· | ···················· | ···················· | ···················· | |
| E | 4944 hCoV-19/USA/CA-ALSR-1066-SAN/2020\|EPI_ISL_483496\|2 | ···················· | ·A·················· | ···················· | ···················· | ···················· | ···················· | |
| E | 4945 hCoV-19/USA/CA-ALSR-1067-SAN/2020\|EPI_ISL_483497\|2 | ···················· | ···················· | ···················· | ···················· | ···················· | ···················· | |
| E | 4946 hCoV-19/USA/CA-ALSR-1068-SAN/2020\|EPI_ISL_483498\|2 | ···················· | ·A·················· | ···················· | ···················· | ···················· | ···················· | |
| E | 4947 hCoV-19/USA/CA-ALSR-1070-SAN/2020\|EPI_ISL_483499\|2 | ···················· | ···················· | ···················· | ···················· | ···················· | ···················· | |
| E | 4951 hCoV-19/USA/MI-UM-2006S001009/2020\|EPI_ISL_484789\| | ···················· | ···················· | ···················· | ···················· | ···················· | ···················· | |
| E | 4953 hCoV-19/USA/MI-UM-2006S001036/2020\|EPI_ISL_484794\| | ···················· | ···················· | ···················· | ···················· | ···················· | ···················· | |
| E | 4954 hCoV-19/USA/MI-UM-2006S001040/2020\|EPI_ISL_484796\| | ···················· | ···················· | ···················· | ···················· | ···················· | ···················· | |
| E | 4955 hCoV-19/England/BRIS-12FAEE/2020\|EPI_ISL_488325\|20 | ···················· | ···················· | ···················· | ···················· | ···················· | ···················· | |
| E | 4956 hCoV-19/England/BRIS-12F36B/2020\|EPI_ISL_488326\|20 | ···················· | ···················· | ···················· | ···················· | ···················· | ···················· | |
| E | 4957 hCoV-19/England/BRIS-12F046/2020\|EPI_ISL_488327\|20 | ···················· | ···················· | ···················· | ···················· | ···················· | ···················· | |
| E | 4958 hCoV-19/England/BRIS-12F5FC/2020\|EPI_ISL_488328\|20 | ···················· | ···················· | ···················· | ···················· | ···················· | ···················· | |
| E | 4959 hCoV-19/England/BRIS-12E5DF/2020\|EPI_ISL_488330\|20 | ···················· | ···················· | ···················· | ···················· | ···················· | ···················· | |
| E | 4960 hCoV-19/England/BRIS-12F486/2020\|EPI_ISL_488331\|20 | ···················· | ···················· | ···················· | ···················· | ···················· | ···················· | |
| E | 4961 hCoV-19/Scotland/GCVR-17141C/2020\|EPI_ISL_489600\|2 | ···················· | ···················· | ···················· | ···················· | ···················· | ···················· | |
| E | 4962 hCoV-19/Scotland/GCVR-170EB5/2020\|EPI_ISL_489601\|2 | ···················· | ···················· | ···················· | ···················· | ···················· | ···················· | |
| E | 4963 hCoV-19/Scotland/GCVR-170F1C/2020\|EPI_ISL_489602\|2 | ···················· | ···················· | ···················· | ···················· | ···················· | ···················· | |
| E | 4964 hCoV-19/Scotland/GCVR-16FBEF/2020\|EPI_ISL_489603\|2 | ···················· | ······A············· | ···················· | ···················· | ···················· | ···················· | |
| E | 4965 hCoV-19/Scotland/GCVR-170F49/2020\|EPI_ISL_489604\|2 | ···················· | ···················· | ···················· | ···················· | ···················· | ···················· | |
| E | 4966 hCoV-19/Scotland/GCVR-171467/2020\|EPI_ISL_489606\|2 | ···················· | ···················· | ···················· | ···················· | ···················· | ···················· | |
| E | 4967 hCoV-19/Scotland/GCVR-16FDDA/2020\|EPI_ISL_489608\|2 | ···················· | ·A·················· | ···················· | ···················· | ···················· | ···················· | |
| E | 4968 hCoV-19/Scotland/GCVR-1717B9/2020\|EPI_ISL_489612\|2 | ···················· | ···················· | ···················· | ···················· | ···················· | ···················· | |
| E | 4969 hCoV-19/Scotland/GCVR-1713D3/2020\|EPI_ISL_489613\|2 | ···················· | ···················· | ···················· | ···················· | ···················· | ···················· | |
| E | 4970 hCoV-19/Scotland/GCVR-1718E3/2020\|EPI_ISL_489615\|2 | ···················· | ···················A | ···················· | ···················· | ···················· | ···················· | 1 |
| E | 4971 hCoV-19/Scotland/GCVR-1716E9/2020\|EPI_ISL_489616\|2 | ···················· | ·A·················· | ···················· | ···················· | ···················· | ···················· | |
| E | 4972 hCoV-19/Scotland/GCVR-1718A7/2020\|EPI_ISL_489617\|2 | ···················· | ···················· | ···················· | ···················· | ···················· | ···················· | |
| E | 4973 hCoV-19/Scotland/GCVR-170E97/2020\|EPI_ISL_489619\|2 | ···················· | ···················· | ···················· | ···················· | ···················· | ···················· | |
| E | 4974 hCoV-19/Scotland/GCVR-17176E/2020\|EPI_ISL_489620\|2 | ···················· | ··············C····· | ···················· | ···················· | ···················· | ···················· | |
| E | 4975 hCoV-19/Scotland/GCVR-16FEAA/2020\|EPI_ISL_489621\|2 | ···················· | ···················· | ···················· | ···················· | ···················· | ···················· | |
| E | 4976 hCoV-19/Scotland/GCVR-1714FE/2020\|EPI_ISL_489622\|2 | ···················· | ···················· | ···················· | ···················· | ···················· | ···················· | |

| | | | | | | | |
|---|---|---|---|---|---|---|---|
| E | 4977 hCoV-19/Scotland/GCVR-171449/2020\|EPI_ISL_489656\|2 | ···················· | ···················· | ···················· | ···················· | ···················· | ···················· |
| E | 4978 hCoV-19/Scotland/GCVR-170811/2020\|EPI_ISL_489657\|2 | ···················· | ···················· | ···················· | ···················· | ···················· | ···················· |
| E | 4979 hCoV-19/Scotland/GCVR-17182F/2020\|EPI_ISL_489658\|2 | ···················· | ···················· | ···················· | ···················· | ···················· | ···················· |
| E | 4980 hCoV-19/Scotland/GCVR-171889/2020\|EPI_ISL_489660\|2 | ···················· | ···················· | ···················· | ···················· | ···················· | ···················· |
| E | 4981 hCoV-19/England/SHEF-C5EB2/2020\|EPI_ISL_490601\|202 | ···················· | ···················· | ···················· | ···················· | ···················· | ···················· |
| E | 4982 hCoV-19/England/SHEF-C6EFD/2020\|EPI_ISL_490605\|202 | ···················· | ···················· | ···················· | ···················· | ···················· | ···················· |
| E | 4983 hCoV-19/England/SHEF-C73DF/2020\|EPI_ISL_490607\|202 | ···················· | ···················· | ···················· | ···················· | ···················· | ···················· |
| E | 4984 hCoV-19/England/SHEF-C7ACB/2020\|EPI_ISL_490608\|202 | ···················· | ···················· | ···················· | ···················· | ···················· | ···················· |
| E | 4985 hCoV-19/England/SHEF-D0E69/2020\|EPI_ISL_490639\|202 | ···················· | ···················· | ···················· | ···················· | ···················· | ···················· |
| E | 4986 hCoV-19/USA/MD_NIDDL_2941/2020\|EPI_ISL_491907\|2020 | ···················· | ···················· | ···················· | ···················· | ···················· | ···················· |
| E | 4987 hCoV-19/USA/MD_NIDDL_2981/2020\|EPI_ISL_491908\|2020 | ···················· | ···················· | ···················· | ···················· | ···················· | ···················· |
| E | 4988 hCoV-19/USA/MD_NIDDL_3202/2020\|EPI_ISL_491909\|2020 | ···················· | ···················· | ···················· | ···················· | ···················· | ···················· |
| E | 4989 hCoV-19/USA/VA_NIDDL_2677/2020\|EPI_ISL_491910\|2020 | ···················· | ···················· | ···················· | ···················· | ···················· | ···················· |
| E | 4990 hCoV-19/USA/VA_NIDDL_2686/2020\|EPI_ISL_491911\|2020 | ···················· | ···················· | ···················· | ···················· | ···················· | ···················· |
| E | 4991 hCoV-19/USA/VA_NIDDL_2689/2020\|EPI_ISL_491912\|2020 | ···················· | ···················· | ···················· | ···················· | ···················· | ···················· |
| E | 4992 hCoV-19/USA/VA_NIDDL_2702/2020\|EPI_ISL_491913\|2020 | ···················· | ···················· | ···················· | ···················· | ···················· | ···················· |
| E | 4993 hCoV-19/USA/VA_NIDDL_2715/2020\|EPI_ISL_491914\|2020 | ···················· | ···················· | ···················· | ···················· | ···················· | ···················· |
| E | 4994 hCoV-19/USA/VA_NIDDL_2716/2020\|EPI_ISL_491915\|2020 | ···················· | ···················· | ···················· | ···················· | ···················· | ···················· |
| E | 4995 hCoV-19/USA/VA_NIDDL_2768/2020\|EPI_ISL_491916\|2020 | ···················· | ···················· | ···················· | ···················· | ···················· | ···················· |
| E | 4996 hCoV-19/USA/VA_NIDDL_2783/2020\|EPI_ISL_491917\|2020 | ···················· | ···················· | ···················· | ···················· | ···················· | ···················· |
| E | 4997 hCoV-19/USA/VA_NIDDL_2791/2020\|EPI_ISL_491918\|2020 | ···················· | ···················· | ···················· | ···················· | ···················· | ···················· |
| E | 4998 hCoV-19/USA/VA_NIDDL_2809/2020\|EPI_ISL_491919\|2020 | ···················· | ···················· | ···················· | ···················· | ···················· | ···················· |
| E | 4999 hCoV-19/USA/VA_NIDDL_2812/2020\|EPI_ISL_491920\|2020 | ···················· | ···················· | ···················· | ···················· | ···················· | ···················· |
| E | 5000 hCoV-19/USA/VA_NIDDL_2896/2020\|EPI_ISL_491921\|2020 | ···················· | ···················· | ···················· | ···················· | ···················· | ···················· |
| E | 5001 hCoV-19/USA/VA_NIDDL_2925/2020\|EPI_ISL_491922\|2020 | ···················· | ···················· | ···················· | ···················· | ···················· | ···················· |
| E | 5002 hCoV-19/USA/VA_NIDDL_2929/2020\|EPI_ISL_491923\|2020 | ···················· | ···················· | ···················· | ···················· | ···················· | ···················· |
| E | 5003 hCoV-19/USA/VA_NIDDL_2932/2020\|EPI_ISL_491924\|2020 | ···················· | ···················· | ···················· | ···················· | ···················· | ···················· |
| E | 5004 hCoV-19/USA/VA_NIDDL_2951/2020\|EPI_ISL_491925\|2020 | ···················· | ···················· | ···················· | ···················· | ···················· | ···················· |
| E | 5005 hCoV-19/USA/VA_NIDDL_2996/2020\|EPI_ISL_491926\|2020 | ···················· | ···················· | ···················· | ···················· | ···················· | ···················· |
| E | 5006 hCoV-19/USA/VA_NIDDL_3033/2020\|EPI_ISL_491927\|2020 | ···················· | ···················· | ···················· | ···················· | ···················· | ···················· |
| E | 5007 hCoV-19/USA/VA_NIDDL_3086/2020\|EPI_ISL_491928\|2020 | ···················· | ···················· | ···················· | ···················· | ···················· | ···················· |
| E | 5008 hCoV-19/USA/VA_NIDDL_3089/2020\|EPI_ISL_491929\|2020 | ···················· | ···················· | ···················· | ···················· | ···················· | ···················· |
| E | 5009 hCoV-19/USA/VA_NIDDL_3090/2020\|EPI_ISL_491930\|2020 | ···················· | ···················· | ···················· | ···················· | ···················· | ···················· |
| E | 5010 hCoV-19/USA/VA_NIDDL_3129/2020\|EPI_ISL_491931\|2020 | ···················· | ···················· | ···················· | ···················· | ···················· | ···················· |
| E | 5011 hCoV-19/Ecuador/USFQ-120/2020\|EPI_ISL_491932\|2020- | ···················· | ···················· | ···················· | ···················· | ···················· | ···················· |
| E | 5012 hCoV-19/Ecuador/USFQ-128/2020\|EPI_ISL_491933\|2020- | ···················· | ···················· | ···················· | ···················· | ···················· | ···················· |
| E | 5013 hCoV-19/Ecuador/USFQ-129/2020\|EPI_ISL_491934\|2020- | ···················· | ···················· | ···················· | ···················· | ···················· | ···················· |
| E | 5014 hCoV-19/Ecuador/USFQ-131/2020\|EPI_ISL_491935\|2020- | ···················· | ···················· | ···················· | ···················· | ···················· | ···················· |
| E | 5015 hCoV-19/Ecuador/USFQ-139/2020\|EPI_ISL_491936\|2020- | ···················· | ···················· | ···················· | ···················· | ···················· | ···················· |
| E | 5016 hCoV-19/Ecuador/USFQ-147/2020\|EPI_ISL_491937\|2020- | ···················· | ···················· | ···················· | ···················· | ···················· | ···················· |
| E | 5017 hCoV-19/Ecuador/USFQ-150/2020\|EPI_ISL_491938\|2020- | ···················· | ···················· | ···················· | ···················· | ···················· | ···················· |
| E | 5018 hCoV-19/Ecuador/USFQ-126/2020\|EPI_ISL_491939\|2020- | ···················· | ···················· | ···················· | ···················· | ···················· | ···················· |
| E | 5020 hCoV-19/Wales/PHWC-166CF4/2020\|EPI_ISL_494104\|2020 | ···················· | ···················· | ···················· | ···················· | ···················· | ···················· |
| E | 5021 hCoV-19/Wales/PHWC-166D00/2020\|EPI_ISL_494105\|2020 | ···················· | ···················· | ···················· | ···················· | ···················· | ···················· |
| E | 5022 hCoV-19/Wales/PHWC-166D2E/2020\|EPI_ISL_494106\|2020 | ···················· | ···················· | ···················· | ···················· | ···················· | ···················· |
| E | 5023 hCoV-19/Wales/PHWC-166D3D/2020\|EPI_ISL_494107\|2020 | ···················· | ···················· | ···················· | ···················· | ···················· | ···················· |
| E | 5024 hCoV-19/Wales/PHWC-166D4C/2020\|EPI_ISL_494108\|2020 | ···················· | ···················· | ···················· | ···················· | ···················· | ···················· |
| E | 5025 hCoV-19/Wales/PHWC-166D6A/2020\|EPI_ISL_494109\|2020 | ···················· | ···················· | ···················· | ···················· | ···················· | ···················· |
| E | 5027 hCoV-19/Wales/PHWC-166E67/2020\|EPI_ISL_494122\|2020 | ···················· | ···················· | ···················· | ···················· | ···················· | ···················· |
| E | 5028 hCoV-19/Wales/PHWC-166E85/2020\|EPI_ISL_494123\|2020 | ···················· | ···················· | ···················· | ···················· | ···················· | ···················· |
| E | 5030 hCoV-19/Wales/PHWC-166EC1/2020\|EPI_ISL_494127\|2020 | ···················· | ···················· | ···················· | ···················· | ···················· | ···················· |
| E | 5031 hCoV-19/Wales/PHWC-166ED0/2020\|EPI_ISL_494128\|2020 | ···················· | ···················· | ···················· | ···················· | ···················· | ···················· |
| E | 5033 hCoV-19/Wales/PHWC-166F0A/2020\|EPI_ISL_494130\|2020 | ···················· | ···················· | ···················· | ···················· | ·NN················· | ···················· |
| E | 5034 hCoV-19/Wales/PHWC-166F55/2020\|EPI_ISL_494132\|2020 | ···················· | ···················· | ···················· | ···················· | ···················· | ···················· |
| E | 5035 hCoV-19/Wales/PHWC-166F64/2020\|EPI_ISL_494133\|2020 | ···················· | ···················· | ···················· | ···················· | ···················· | ···················· |
| E | 5036 hCoV-19/Wales/PHWC-166F82/2020\|EPI_ISL_494134\|2020 | ···················· | ···················· | ···················· | ···················· | ···················· | ···················· |
| E | 5037 hCoV-19/Wales/PHWC-166FA0/2020\|EPI_ISL_494135\|2020 | ···················· | ···················· | ···················· | ···················· | ···················· | ···················· |
| E | 5040 hCoV-19/Wales/PHWC-167033/2020\|EPI_ISL_494143\|2020 | ···················· | ···················· | ···················· | ···················· | ···················· | ···················· |
| E | 5041 hCoV-19/Wales/PHWC-167042/2020\|EPI_ISL_494144\|2020 | ···················· | ···················· | ···················· | ···················· | ···················· | ···················· |
| E | 5042 hCoV-19/Wales/PHWC-167060/2020\|EPI_ISL_494146\|2020 | ···················· | ···················· | ···················· | ···················· | ···················· | ···················· |
| E | 5043 hCoV-19/Wales/PHWC-167DA5/2020\|EPI_ISL_494156\|2020 | ···················· | ···················· | ···················· | ···················· | ···················· | ···················· |
| E | 5044 hCoV-19/Wales/PHWC-167DE1/2020\|EPI_ISL_494159\|2020 | ···················· | ···················· | ···················· | ···················· | ···················· | ···················· |
| E | 5045 hCoV-19/Wales/PHWC-167E0C/2020\|EPI_ISL_494161\|2020 | ···················· | ···················· | ···················· | ···················· | ···················· | ···················· |
| E | 5046 hCoV-19/Wales/PHWC-167E39/2020\|EPI_ISL_494163\|2020 | ···················· | ···················· | ···················· | ···················· | ···················· | ···················· |
| E | 5047 hCoV-19/Wales/PHWC-167E57/2020\|EPI_ISL_494165\|2020 | ···················· | ···················· | ···················· | ···················· | ···················· | ···················· |
| E | 5048 hCoV-19/Wales/PHWC-167E84/2020\|EPI_ISL_494167\|2020 | ···················· | ···················· | ···················· | ···················· | ···················· | ···················· |
| E | 5049 hCoV-19/Wales/PHWC-167E93/2020\|EPI_ISL_494168\|2020 | ···················· | ···················· | ···················· | ···················· | ···················· | ···················· |
| E | 5050 hCoV-19/Wales/PHWC-167EEE/2020\|EPI_ISL_494173\|2020 | ···················· | ···················· | ···················· | ···················· | ···················· | ···················· |
| E | 5051 hCoV-19/Wales/PHWC-167EFD/2020\|EPI_ISL_494174\|2020 | ···················· | ···················· | ···················· | ···················· | ···················· | ···················· |
| E | 5052 hCoV-19/Wales/PHWC-167F09/2020\|EPI_ISL_494175\|2020 | ···················· | ···················· | ···················· | ···················· | ···················· | ···················· |
| E | 5053 hCoV-19/Wales/PHWC-167F27/2020\|EPI_ISL_494176\|2020 | ···················· | ···················· | ···················· | ···················· | ···················· | ···················· |
| E | 5054 hCoV-19/Wales/PHWC-167F45/2020\|EPI_ISL_494178\|2020 | ···················· | ···················· | ···················· | ···················· | ···················· | ···················· |
| E | 5055 hCoV-19/Wales/PHWC-167F63/2020\|EPI_ISL_494179\|2020 | ···················· | ···················· | ···················· | ···················· | ···················· | ···················· |
| E | 5056 hCoV-19/Wales/PHWC-167F81/2020\|EPI_ISL_494181\|2020 | ···················· | ···················· | ···················· | ···················· | ···················· | ···················· |
| E | 5057 hCoV-19/Wales/PHWC-167FCD/2020\|EPI_ISL_494185\|2020 | ···················· | ···················· | ···················· | ···················· | ···················· | ···················· |
| E | 5058 hCoV-19/Wales/PHWC-167FEB/2020\|EPI_ISL_494187\|2020 | ···················· | ···················· | ···················· | ···················· | ···················· | ···················· |
| E | 5059 hCoV-19/Wales/PHWC-16807E/2020\|EPI_ISL_494195\|2020 | ···················· | ···················· | ···················· | ···················· | ···················· | ···················· |
| E | 5060 hCoV-19/Turkey/KU-010/2020\|EPI_ISL_495420\|2020-05- | ···················· | ···················· | ···················· | ···················· | ···················· | ···················· |
| E | 5061 hCoV-19/Turkey/KU-011/2020\|EPI_ISL_495421\|2020-06- | ···················· | ···················· | ···················· | ···················· | ···················· | ···················· |
| E | 5062 hCoV-19/Turkey/KU-012/2020\|EPI_ISL_495422\|2020-05- | ···················· | ···················· | ···················· | ···················· | ···················· | ···················· |

| | | | | | | | | |
|---|---|---|---|---|---|---|---|---|
| E | 5063 | hCoV-19/Turkey/KU-013/2020\|EPI_ISL_495423\|2020-05- | ···················· | ···················· | ···················· | ···················· | ···················· | ···················· |
| E | 5064 | hCoV-19/Turkey/KU-015/2020\|EPI_ISL_495425\|2020-05- | ···················· | ···················· | ···················· | ···················· | ···················· | ···················· |
| E | 5065 | hCoV-19/Turkey/KU-016/2020\|EPI_ISL_495426\|2020-05- | ···················· | ···················· | ···················· | ···················· | ···················· | ···················· |
| E | 5066 | hCoV-19/Turkey/KU-018/2020\|EPI_ISL_495428\|2020-04- | ···················· | ···················· | ···················· | ···················· | ···················· | ···················· |
| E | 5067 | hCoV-19/Turkey/KU-019/2020\|EPI_ISL_495429\|2020-06- | ···················· | ···················· | ···················· | ···················· | ···················· | ···················· |
| E | 5068 | hCoV-19/Turkey/KU-020/2020\|EPI_ISL_495430\|2020-05- | ···················· | ···················· | ···················· | ···················· | ···················· | ···················· |
| E | 5069 | hCoV-19/Turkey/KU-021/2020\|EPI_ISL_495431\|2020-05- | ···················· | ···················· | ···················· | ···················· | ···················· | ···················· |
| E | 5070 | hCoV-19/Turkey/KU-022/2020\|EPI_ISL_495432\|2020-04- | ···················· | ···················· | ···················· | ···················· | ···················· | ···················· |
| E | 5071 | hCoV-19/Turkey/KU-023/2020\|EPI_ISL_495433\|2020-06- | ···················· | ···················· | ···················· | ···················· | ···················· | ···················· |
| E | 5072 | hCoV-19/Turkey/KU-024/2020\|EPI_ISL_495434\|2020-05- | ···················· | ···················· | ···················· | ···················· | ···················· | ···················· |
| E | 5073 | hCoV-19/Turkey/KU-025/2020\|EPI_ISL_495435\|2020-05- | ···················· | ···················· | ···················· | ···················· | ···················· | ···················· |
| E | 5074 | hCoV-19/Turkey/KU-026/2020\|EPI_ISL_495436\|2020-06- | ···················· | ···················· | ···················· | ···················· | ···················· | ···················· |
| E | 5075 | hCoV-19/Turkey/KU-027/2020\|EPI_ISL_495437\|2020-05- | ···················· | ···················· | ···················· | ···················· | ···················· | ···················· |
| E | 5076 | hCoV-19/Turkey/KU-028/2020\|EPI_ISL_495438\|2020-05- | ···················· | ···················· | ···················· | ···················· | ···················· | ···················· |
| E | 5077 | hCoV-19/Turkey/KU-029/2020\|EPI_ISL_495439\|2020-05- | ···················· | ···················· | ···················· | ···················· | ···················· | ···················· |
| E | 5078 | hCoV-19/Turkey/KU-031/2020\|EPI_ISL_495441\|2020-04- | ···················· | ···················· | ···················· | ···················· | ···················· | ···················· |
| E | 5079 | hCoV-19/Turkey/KU-034/2020\|EPI_ISL_495444\|2020-05- | ···················· | ···················· | ···················· | ···················· | ···················· | ···················· |
| E | 5080 | hCoV-19/Turkey/KU-043/2020\|EPI_ISL_495453\|2020-05- | ···················· | ···················· | ···················· | ···················· | ···················· | ···················· |
| E | 5081 | hCoV-19/Turkey/KU-044/2020\|EPI_ISL_495454\|2020-05- | ···················· | ···················· | ···················· | ···················· | ···················· | ···················· |
| E | 5082 | hCoV-19/Turkey/KU-045/2020\|EPI_ISL_495455\|2020-05- | ···················· | ···················· | ···················· | ···················· | ···················· | ···················· |
| E | 5083 | hCoV-19/Turkey/KU-046/2020\|EPI_ISL_495456\|2020-05- | ···················· | ···················· | ···················· | ···················· | ···················· | ···················· |
| E | 5084 | hCoV-19/Turkey/KU-047/2020\|EPI_ISL_495457\|2020-05- | ···················· | ···················· | ···················· | ···················· | ···················· | ···················· |
| E | 5085 | hCoV-19/Lishui/LS003/2020\|EPI_ISL_495459\|2020-01-2 | ···················· | ···················· | ···················· | ···················· | ···················· | ···················· |
| E | 5086 | hCoV-19/USA/NJ-UW-755/2020\|EPI_ISL_495460\|2020-07- | ···················· | ···················· | ···················· | ···················· | ···················· | ···················· |
| E | 5087 | hCoV-19/USA/WI-UW-756/2020\|EPI_ISL_495461\|2020-07- | ···················· | ···················· | ···················· | ···················· | ···················· | ···················· |
| E | 5088 | hCoV-19/USA/WI-UW-757/2020\|EPI_ISL_495462\|2020-07- | ···················· | ···················· | ···················· | ···················· | ···················· | ···················· |
| E | 5089 | hCoV-19/USA/WI-UW-758/2020\|EPI_ISL_495463\|2020-07- | ···················· | ···················· | ···················· | ···················· | ···················· | ···················· |
| E | 5090 | hCoV-19/USA/WI-UW-760/2020\|EPI_ISL_495465\|2020-07- | ···················· | ···················· | ···················· | ···················· | ···················· | ···················· |
| E | 5091 | hCoV-19/USA/WI-UW-762/2020\|EPI_ISL_495467\|2020-07- | ···················· | ···················· | ···················· | ···················· | ···················· | ···················· |
| E | 5092 | hCoV-19/USA/WI-UW-763/2020\|EPI_ISL_495468\|2020-07- | ···················· | ···················· | ···················· | ···················· | ···················· | ···················· |
| E | 5093 | hCoV-19/USA/WI-UW-765/2020\|EPI_ISL_495470\|2020-07- | ···················· | ···················· | ···················· | ···················· | ···················· | ···················· |
| E | 5094 | hCoV-19/USA/WI-UW-766/2020\|EPI_ISL_495471\|2020-07- | ···················· | ···················· | ···················· | ···················· | ···················· | ···················· |
| E | 5095 | hCoV-19/USA/WI-UW-767/2020\|EPI_ISL_495472\|2020-07- | ···················· | ···················· | ···················· | ···················· | ···················· | ···················· |
| E | 5096 | hCoV-19/USA/WI-UW-768/2020\|EPI_ISL_495473\|2020-07- | ···················· | ···················· | ···················· | ···················· | ···················· | ···················· |
| E | 5097 | hCoV-19/USA/WI-UW-769/2020\|EPI_ISL_495474\|2020-07- | ···················· | ···················· | ···················· | ···················· | ···················· | ···················· |
| E | 5098 | hCoV-19/USA/WI-UW-770/2020\|EPI_ISL_495475\|2020-07- | ···················· | ···················· | ···················· | ···················· | ···················· | ···················· |
| E | 5099 | hCoV-19/USA/MO-UW-771/2020\|EPI_ISL_495476\|2020-07- | ············T······· | ···················· | ···················· | ···················· | ···················· | ···················· |
| E | 5102 | hCoV-19/USA/WI-UW-776/2020\|EPI_ISL_495480\|2020-07- | ···················· | ···················· | ···················· | ···················· | ···················· | ···················· |
| E | 5103 | hCoV-19/USA/WI-UW-777/2020\|EPI_ISL_495481\|2020-07- | ···················· | ···················· | ···················· | ···················· | ···················· | ···················· |
| E | 5104 | hCoV-19/USA/WI-UW-778/2020\|EPI_ISL_495482\|2020-07- | ···················· | ···················· | ···················· | ···················· | ···················· | ···················· |
| E | 5105 | hCoV-19/USA/WI-UW-779/2020\|EPI_ISL_495483\|2020-07- | ···················· | ···················· | ···················· | ···················· | ···················· | ···················· |
| E | 5106 | hCoV-19/USA/WI-UW-780/2020\|EPI_ISL_495484\|2020-07- | ···················· | ···················· | ···················· | ···················· | ···················· | ···················· |
| E | 5107 | hCoV-19/USA/WI-UW-781/2020\|EPI_ISL_495485\|2020-07- | ···················· | ···················· | ···················· | ···················· | ···················· | ···················· |
| E | 5108 | hCoV-19/USA/WI-UW-782/2020\|EPI_ISL_495486\|2020-07- | ···················· | ···················· | ···················· | ···················· | ···················· | ···················· |
| E | 5109 | hCoV-19/USA/WI-UW-783/2020\|EPI_ISL_495487\|2020-07- | ···················· | ···················· | ···················· | ···················· | ···················· | ···················· |
| E | 5110 | hCoV-19/USA/WI-UW-785/2020\|EPI_ISL_495489\|2020-07- | ···················· | ···················· | ···················· | ···················· | ···················· | ···················· |
| E | 5111 | hCoV-19/USA/WI-UW-786/2020\|EPI_ISL_495490\|2020-07- | ···················· | ···················· | ···················· | ···················· | ···················· | ···················· |
| E | 5113 | hCoV-19/USA/WI-UW-788/2020\|EPI_ISL_495492\|2020-07- | ···················· | ···················· | ···················· | ···················· | ···················· | ···················· |
| E | 5114 | hCoV-19/USA/WI-UW-789/2020\|EPI_ISL_495493\|2020-07- | ···················· | ···················· | ···················· | ···················· | ···················· | ···················· |
| E | 5115 | hCoV-19/USA/WI-UW-790/2020\|EPI_ISL_495494\|2020-07- | ···················· | ···················· | ···················· | ···················· | ···················· | ···················· |
| E | 5116 | hCoV-19/USA/WI-UW-791/2020\|EPI_ISL_495495\|2020-07- | ···················· | ···················· | ···················· | ···················· | ···················· | ···················· |
| E | 5117 | hCoV-19/USA/WI-UW-792/2020\|EPI_ISL_495496\|2020-07- | ···················· | ···················· | ···················· | ···················· | ···················· | ···················· |
| E | 5118 | hCoV-19/USA/WI-UW-793/2020\|EPI_ISL_495497\|2020-07- | ···················· | ···················· | ···················· | ···················· | ···················· | ···················· |
| E | 5119 | hCoV-19/USA/WI-UW-794/2020\|EPI_ISL_495498\|2020-07- | ···················· | ···················· | ···················· | ···················· | ···················· | ···················· |
| E | 5120 | hCoV-19/USA/WI-UW-795/2020\|EPI_ISL_495499\|2020-07- | ···················· | ···················· | ···················· | ···················· | ···················· | ···················· |
| E | 5121 | hCoV-19/England/HECH-2599E8F/2020\|EPI_ISL_482100\|2 | ···················· | ···················· | ···················· | ···················· | ···················· | ···················· |
| E | 5122 | hCoV-19/Northern Ireland/NIRE-101402/2020\|EPI_ISL_ | ···················· | ···················· | ···················· | ···················· | ···················· | ···················· |
| E | 5123 | hCoV-19/Northern Ireland/NIRE-10289C/2020\|EPI_ISL_ | ···················· | ···················· | ···················· | ···················· | ···················· | ···················· |
| E | 5124 | hCoV-19/England/HECH-2599ECB/2020\|EPI_ISL_482103\|2 | ···················· | ···················· | ···················· | ···················· | ···················· | ···················· |
| E | 5125 | hCoV-19/England/HECH-2599DBF/2020\|EPI_ISL_482104\|2 | ···················· | ···················· | ···················· | ···················· | ···················· | ···················· |
| E | 5126 | hCoV-19/Northern Ireland/NIRE-FF7C3/2020\|EPI_ISL_4 | ···················· | ···················· | ···················· | ···················· | ···················· | ···················· |
| E | 5127 | hCoV-19/England/HECH-2599FE6/2020\|EPI_ISL_482106\|2 | ···················· | ···················· | ···················· | ···················· | ···················· | ···················· |
| E | 5128 | hCoV-19/England/HECH-2599DCE/2020\|EPI_ISL_482108\|2 | ···················· | ···················· | ···················· | ···················· | ···················· | ···················· |
| E | 5129 | hCoV-19/England/HECH-2599C58/2020\|EPI_ISL_482109\|2 | ···················· | ···················· | ···················· | ···················· | ···················· | ···················· |
| E | 5130 | hCoV-19/Northern Ireland/NIRE-100F7B/2020\|EPI_ISL_ | ···················· | ···················· | ···················· | ···················· | ···················· | ···················· |
| E | 5131 | hCoV-19/Northern Ireland/NIRE-100DAE/2020\|EPI_ISL_ | ···················· | ···················· | ···················· | ···················· | ···················· | ···················· |
| E | 5132 | hCoV-19/Northern Ireland/NIRE-100B4A/2020\|EPI_ISL_ | ···················· | ···················· | ···················· | ···················· | ···················· | ···················· |
| E | 5133 | hCoV-19/Northern Ireland/NIRE-10100E/2020\|EPI_ISL_ | ···················· | ···················· | ···················· | ···················· | ···················· | ···················· |
| E | 5134 | hCoV-19/USA/CA-ALSR-0899-IPL/2020\|EPI_ISL_483403\|2 | ···················· | ·A·················· | ···················· | ···················· | ···················· | ···················· |
| E | 5135 | hCoV-19/USA/CA-ALSR-0901-SAN/2020\|EPI_ISL_483404\|2 | ···················· | ·A·················· | ···················· | ···················· | ···················· | ···················· |
| E | 5136 | hCoV-19/USA/CA-ALSR-0902-IPL/2020\|EPI_ISL_483405\|2 | ···················· | ·A·················· | ···················· | ···················· | ···················· | ···················· |
| E | 5137 | hCoV-19/USA/CA-ALSR-0903-IPL/2020\|EPI_ISL_483406\|2 | ···················· | ·A·················· | ···················· | ···················· | ···················· | ···················· |
| E | 5138 | hCoV-19/USA/CA-ALSR-0905-IPL/2020\|EPI_ISL_483407\|2 | ···················· | ···················· | ···················· | ···················· | ···················· | ···················· |
| E | 5139 | hCoV-19/USA/CA-ALSR-0907-IPL/2020\|EPI_ISL_483409\|2 | ···················· | ·A·················· | ···················· | ···················· | ···················· | ···················· |
| E | 5140 | hCoV-19/USA/CA-ALSR-0910-SAN/2020\|EPI_ISL_483410\|2 | ···················· | ·A·················· | ···················· | ···················· | ···················· | ···················· |
| E | 5141 | hCoV-19/USA/CA-ALSR-0918-IPL/2020\|EPI_ISL_483413\|2 | ···················· | ·A·················· | ···················· | ···················· | ···················· | ···················· |
| E | 5142 | hCoV-19/USA/CA-ALSR-0921-IPL/2020\|EPI_ISL_483414\|2 | ···················· | ·A·················· | ···················· | ···················· | ···················· | ···················· |
| E | 5143 | hCoV-19/USA/CA-ALSR-0923-IPL/2020\|EPI_ISL_483415\|2 | ···················· | ·A·················· | ···················· | ···················· | ···················· | ···················· |
| E | 5144 | hCoV-19/USA/CA-ALSR-0925-IPL/2020\|EPI_ISL_483416\|2 | ···················· | ·A·················· | ···················· | ···················· | ···················· | ···················· |
| E | 5145 | hCoV-19/USA/CA-ALSR-0926-IPL/2020\|EPI_ISL_483417\|2 | ···················· | ·A·················· | ···················· | ···················· | ···················· | ···················· |

| | | | | | | | | |
|---|---|---|---|---|---|---|---|---|
| E | 5146 | hCoV-19/USA/CA-ALSR-0928-SAN/2020\|EPI_ISL_483418\|2 | ···················· | ·A·················· | ···················· | ···················· | ···················· | ···················· |
| E | 5147 | hCoV-19/USA/CA-ALSR-0930-IPL/2020\|EPI_ISL_483419\|2 | ···················· | ···················· | ···················· | ···················· | ···················· | ···················· |
| E | 5148 | hCoV-19/USA/CA-ALSR-0932-IPL/2020\|EPI_ISL_483421\|2 | ···················· | ···················· | ···················· | ···················· | ···················· | ···················· |
| E | 5149 | hCoV-19/USA/CA-ALSR-0934-SAN/2020\|EPI_ISL_483423\|2 | ···················· | ·A·················· | ···················· | ···················· | ···················· | ···················· |
| E | 5150 | hCoV-19/USA/CA-ALSR-0939-SAN/2020\|EPI_ISL_483425\|2 | ···················· | ···················· | ···················· | ···················· | ···················· | ···················· |
| E | 5151 | hCoV-19/USA/CA-ALSR-0941-SAN/2020\|EPI_ISL_483426\|2 | ···················· | ···················· | ···················· | ···················· | ···················· | ···················· |
| E | 5152 | hCoV-19/USA/CA-ALSR-0943-SAN/2020\|EPI_ISL_483427\|2 | ···················· | ···················· | ···················· | ···················· | ···················· | ···················· |
| E | 5153 | hCoV-19/USA/CA-ALSR-0944-SAN/2020\|EPI_ISL_483428\|2 | ···················· | ···················· | ···················· | ···················· | ···················· | ···················· |
| E | 5154 | hCoV-19/USA/CA-ALSR-0945-SAN/2020\|EPI_ISL_483429\|2 | ···················· | ···················· | ···················· | ···················· | ···················· | ···················· |
| E | 5155 | hCoV-19/USA/CA-ALSR-0953-SAN/2020\|EPI_ISL_483430\|2 | ···················· | ···················· | ···················· | ···················· | ···················· | ···················· |
| E | 5156 | hCoV-19/USA/CA-ALSR-0955-SAN/2020\|EPI_ISL_483431\|2 | ···················· | ···················· | ···················· | ···················· | ···················· | ···················· |
| E | 5157 | hCoV-19/USA/CA-ALSR-0956-SAN/2020\|EPI_ISL_483432\|2 | ···················· | ···················· | ···················· | ···················· | ···················· | ···················· |
| E | 5158 | hCoV-19/USA/CA-ALSR-0974-IPL/2020\|EPI_ISL_483433\|2 | ···················· | ···················· | ···················· | ···················· | ···················· | ···················· |
| E | 5159 | hCoV-19/USA/CA-ALSR-0975-SAN/2020\|EPI_ISL_483434\|2 | ···················· | ···················· | ···················· | ···················· | ···················· | ···················· |
| E | 5160 | hCoV-19/USA/CA-ALSR-0976-SAN/2020\|EPI_ISL_483435\|2 | ···················· | ···················· | ···················· | ···················· | ···················· | ···················· |
| E | 5161 | hCoV-19/USA/CA-ALSR-0977-SAN/2020\|EPI_ISL_483436\|2 | ···················· | ···················· | ···················· | ···················· | ···················· | ···················· |
| E | 5162 | hCoV-19/USA/CA-ALSR-0979-SAN/2020\|EPI_ISL_483438\|2 | ···················· | ···················· | ···················· | ···················· | ···················· | ···················· |
| E | 5163 | hCoV-19/USA/CA-ALSR-0980-SAN/2020\|EPI_ISL_483439\|2 | ···················· | ···················· | ···················· | ···················· | ···················· | ···················· |
| E | 5164 | hCoV-19/USA/CA-ALSR-0981-SAN/2020\|EPI_ISL_483440\|2 | ···················· | ·A·················· | ···················· | ···················· | ···················· | ···················· |
| E | 5165 | hCoV-19/USA/CA-ALSR-1032-IPL/2020\|EPI_ISL_483474\|2 | ···················· | ·A·················· | ···················· | ···················· | ···················· | ···················· |
| E | 5166 | hCoV-19/USA/CA-ALSR-1035-IPL/2020\|EPI_ISL_483475\|2 | ···················· | ···················· | ···················· | ···················· | ···················· | ···················· |
| E | 5167 | hCoV-19/USA/CA-ALSR-1036-SAN/2020\|EPI_ISL_483476\|2 | ···················· | ·A·················· | ···················· | ···················· | ···················· | ···················· |
| E | 5168 | hCoV-19/USA/CA-ALSR-1038-SAN/2020\|EPI_ISL_483477\|2 | ···················· | ·A·················· | ···················· | ···················· | ···················· | ···················· |
| E | 5169 | hCoV-19/USA/CA-ALSR-1039-SAN/2020\|EPI_ISL_483478\|2 | ···················· | ···················· | ···················· | ···················· | ···················· | ···················· |
| E | 5170 | hCoV-19/USA/CA-ALSR-1042-SAN/2020\|EPI_ISL_483479\|2 | ···················· | ·A·················· | ···················· | ···················· | ···················· | ···················· |
| E | 5171 | hCoV-19/USA/CA-ALSR-1044-SAN/2020\|EPI_ISL_483480\|2 | ···················· | ·A·················· | ···················· | ···················· | ···················· | ···················· |
| E | 5172 | hCoV-19/USA/CA-ALSR-1045-SAN/2020\|EPI_ISL_483481\|2 | ···················· | ·A·················· | ···················· | ···················· | ···················· | ···················· |
| E | 5173 | hCoV-19/USA/CA-ALSR-1049-SAN/2020\|EPI_ISL_483482\|2 | ···················· | ···················· | ···················· | ···················· | ···················· | ···················· |
| E | 5174 | hCoV-19/USA/CA-ALSR-1050-SAN/2020\|EPI_ISL_483483\|2 | ···················· | ···················· | ···················· | ···················· | ···················· | ···················· |
| E | 5175 | hCoV-19/USA/CA-ALSR-1052-SAN/2020\|EPI_ISL_483484\|2 | ···················· | ·A·················· | ···················· | ···················· | ···················· | ···················· |
| E | 5176 | hCoV-19/Belgium/ULG-10212/2020\|EPI_ISL_484700\|2020 | ···················· | ···················· | ···················· | ···················· | ···················· | ···················· |
| E | 5177 | hCoV-19/Belgium/ULG-10214/2020\|EPI_ISL_484702\|2020 | ···················· | ···················· | ···················· | ···················· | ···················· | ···················· |
| E | 5178 | hCoV-19/Belgium/ULG-10215/2020\|EPI_ISL_484703\|2020 | ···················· | ···················· | ···················· | ···················· | ···················· | ···················· |
| E | 5179 | hCoV-19/USA/MI-UM-2006S000850/2020\|EPI_ISL_484709\| | ···················· | ···················· | ···················· | ···················· | ···················· | ···················· |
| E | 5180 | hCoV-19/USA/MI-UM-2006S000851/2020\|EPI_ISL_484710\| | ···················· | ···················· | ···················· | ···················· | ···················· | ···················· |
| E | 5181 | hCoV-19/USA/MI-UM-2006S000852/2020\|EPI_ISL_484711\| | ···················· | ···················· | ···················· | ···················· | ···················· | ···················· |
| E | 5182 | hCoV-19/USA/MI-UM-2006S000853/2020\|EPI_ISL_484712\| | ···················· | ···················· | ···················· | ···················· | ···················· | ···················· |
| E | 5183 | hCoV-19/USA/MI-UM-2006S000856/2020\|EPI_ISL_484713\| | ···················· | ···················· | ···················· | ···················· | ···················· | ···················· |
| E | 5184 | hCoV-19/USA/MI-UM-2006S000857/2020\|EPI_ISL_484714\| | ···················· | ···················· | ···················· | ···················· | ···················· | ···················· |
| E | 5185 | hCoV-19/USA/MI-UM-2006S000858/2020\|EPI_ISL_484715\| | ···················· | ···················· | ···················· | ···················· | ···················· | ···················· |
| E | 5186 | hCoV-19/USA/MI-UM-2006S000863/2020\|EPI_ISL_484718\| | ···················· | ···················· | ···················· | ···················· | ···················· | ···················· |
| E | 5190 | hCoV-19/USA/MI-UM-2006S000870/2020\|EPI_ISL_484723\| | ···················· | ···················· | ···················· | ···················· | ···················· | ···················· |
| E | 5191 | hCoV-19/USA/MI-UM-2006S000872/2020\|EPI_ISL_484724\| | ···················· | ···················· | ···················· | ···················· | ···················· | ···················· |
| E | 5193 | hCoV-19/USA/MI-UM-2006S000881/2020\|EPI_ISL_484731\| | ···················· | ···················· | ···················· | ···················· | ···················· | ···················· |
| E | 5195 | hCoV-19/USA/MI-UM-2006S000883/2020\|EPI_ISL_484733\| | ···················· | ···················· | ···················· | ···················· | ···················· | ···················· |
| E | 5197 | hCoV-19/USA/MI-UM-2006S000915/2020\|EPI_ISL_484753\| | ···················· | ···················· | ···················· | ···················· | ···················· | ···················· |
| E | 5203 | hCoV-19/USA/MI-UM-2006S000934/2020\|EPI_ISL_484762\| | ···················· | ···················· | ···················· | ···················· | ···················· | ···················· |
| E | 5204 | hCoV-19/USA/MI-UM-2006S000935/2020\|EPI_ISL_484763\| | ···················· | ···················· | ···················· | ···················· | ···················· | ···················· |
| E | 5205 | hCoV-19/USA/MI-UM-2006S000944/2020\|EPI_ISL_484768\| | ···················· | ···················· | ···················· | ···················· | ···················· | ···················· |
| E | 5207 | hCoV-19/USA/MI-UM-2006S000960/2020\|EPI_ISL_484770\| | ···················· | ···················· | ···················· | ···················· | ···················· | ···················· |
| E | 5208 | hCoV-19/England/BRIS-12EBEC/2020\|EPI_ISL_488259\|20 | ···················· | ···················· | ···················· | ···················· | ···················· | ···················· |
| E | 5209 | hCoV-19/England/BRIS-12FAA2/2020\|EPI_ISL_488261\|20 | ···················· | ···················· | ···················· | ···················· | ···················· | ···················· |
| E | 5210 | hCoV-19/England/BRIS-12F6CC/2020\|EPI_ISL_488262\|20 | ···················· | ···················· | ···················· | ···················· | ···················· | ···················· |
| E | 5211 | hCoV-19/England/BRIS-12ED7D/2020\|EPI_ISL_488263\|20 | ···················· | ···················· | ···················· | ···················· | ···················· | ···················· |
| E | 5212 | hCoV-19/England/BRIS-12E830/2020\|EPI_ISL_488264\|20 | ···················· | ···················· | ···················· | ···················· | ···················· | ···················· |
| E | 5213 | hCoV-19/England/BRIS-12F4D1/2020\|EPI_ISL_488265\|20 | ···················· | ···················· | ···················· | A··················· | ···················· | ···················· |
| E | 5214 | hCoV-19/England/BRIS-12F35C/2020\|EPI_ISL_488266\|20 | ···················· | ···················· | ···················· | ···················· | ···················· | ···················· |
| E | 5215 | hCoV-19/England/BRIS-12FB63/2020\|EPI_ISL_488267\|20 | ···················· | ···················· | ···················· | ···················· | ···················· | ···················· |
| E | 5216 | hCoV-19/England/BRIS-12F9C3/2020\|EPI_ISL_488268\|20 | ···················· | ···················· | ···················· | ···················· | ···················· | ···················· |
| E | 5217 | hCoV-19/England/BRIS-12F125/2020\|EPI_ISL_488269\|20 | ···················· | ···················· | ···················· | ···················· | ···················· | ···················· |
| E | 5218 | hCoV-19/England/BRIS-12EF68/2020\|EPI_ISL_488271\|20 | ···················· | ···················· | ···················· | ···················· | ···················· | ···················· |
| E | 5219 | hCoV-19/England/BRIS-12F88A/2020\|EPI_ISL_488272\|20 | ···················· | ···················· | ···················· | ···················· | ···················· | ···················· |
| E | 5220 | hCoV-19/England/BRIS-12F7BA/2020\|EPI_ISL_488273\|20 | ···················· | ···················· | ···················· | ···················· | ···················· | ···················· |
| E | 5221 | hCoV-19/England/BRIS-12E672/2020\|EPI_ISL_488274\|20 | ···················· | ···················· | ···················· | ···················· | ···················· | ···················· |
| E | 5222 | hCoV-19/England/BRIS-12E77F/2020\|EPI_ISL_488275\|20 | ···················· | ···················· | ···················· | ···················· | ···················· | ···················· |
| E | 5223 | hCoV-19/England/BRIS-12E733/2020\|EPI_ISL_488276\|20 | ···················· | ···················· | ···················· | ···················· | ···················· | ···················· |
| E | 5224 | hCoV-19/England/BRIS-12F7AB/2020\|EPI_ISL_488277\|20 | ···················· | ···················· | ···················· | ···················· | ···················· | ···················· |
| E | 5225 | hCoV-19/England/BRIS-12FAB1/2020\|EPI_ISL_488280\|20 | ···················· | ···················· | ···················· | ···················· | ···················· | ···················· |
| E | 5226 | hCoV-19/England/BRIS-12E86D/2020\|EPI_ISL_488283\|20 | ···················· | ···················· | ···················· | ···················· | ···················· | ···················· |
| E | 5227 | hCoV-19/England/BRIS-12EC70/2020\|EPI_ISL_488284\|20 | ···················· | ···················· | ···················· | ···················· | ···················· | ···················· |
| E | 5228 | hCoV-19/England/BRIS-12EE5C/2020\|EPI_ISL_488285\|20 | ···················· | ···················· | ···················· | ···················· | ···················· | ···················· |
| E | 5229 | hCoV-19/England/BRIS-12E760/2020\|EPI_ISL_488286\|20 | ···················· | ···················· | ···················· | ···················· | ···················· | ···················· |
| E | 5230 | hCoV-19/England/BRIS-12F723/2020\|EPI_ISL_488287\|20 | ···················· | ···················· | ···················· | ···················· | ···················· | ···················· |
| E | 5231 | hCoV-19/England/BRIS-12E85E/2020\|EPI_ISL_488290\|20 | ···················· | ···················· | ···················· | ···················· | ···················· | ···················· |
| E | 5232 | hCoV-19/England/BRIS-12F3A7/2020\|EPI_ISL_488291\|20 | ···················· | ···················· | ···················· | ···················· | ···················· | ···················· |
| E | 5233 | hCoV-19/England/BRIS-12ECF8/2020\|EPI_ISL_488292\|20 | ···················· | ···················· | ···················· | ···················· | ···················· | ···················· |
| E | 5234 | hCoV-19/England/BRIS-12FA0C/2020\|EPI_ISL_488293\|20 | ···················· | ···················· | ···················· | ···················· | ···················· | ···················· |
| E | 5235 | hCoV-19/England/BRIS-12F2D7/2020\|EPI_ISL_488295\|20 | ···················· | ···················· | ···················· | ···················· | ···················· | ···················· |
| E | 5236 | hCoV-19/England/BRIS-12F28C/2020\|EPI_ISL_488296\|20 | ···················· | ···················· | ···················· | ···················· | ···················· | ···················· |
| E | 5237 | hCoV-19/England/BRIS-12E9E2/2020\|EPI_ISL_488297\|20 | ···················· | ···················· | ···················· | ···················· | ···················· | ···················· |

| | | | | | | | | |
|---|---|---|---|---|---|---|---|---|
| E | 5238 hCoV-19/England/BRIS-12F2C8/2020\|EPI_ISL_488298\|20 | .................... | .................... | .................... | .................... | .................... | .................... | |
| E | 5239 hCoV-19/England/BRIS-12EC8F/2020\|EPI_ISL_488299\|20 | .................... | .................... | .................... | .................... | .................... | .................... | |
| E | 5240 hCoV-19/England/BRIS-12F6F9/2020\|EPI_ISL_488413\|20 | .................... | .................... | .................... | .................... | .................... | .................... | |
| E | 5241 hCoV-19/England/BRIS-12EB73/2020\|EPI_ISL_488415\|20 | .................... | .................... | .................... | .................... | .................... | .................... | |
| E | 5242 hCoV-19/England/BRIS-12F42C/2020\|EPI_ISL_488416\|20 | .................... | .................... | .................... | .................... | .................... | .................... | |
| E | 5243 hCoV-19/England/BRIS-12F41D/2020\|EPI_ISL_488417\|20 | .................... | .................... | .................... | .................... | .................... | .................... | |
| E | 5244 hCoV-19/England/BRIS-12E663/2020\|EPI_ISL_488418\|20 | .................... | .................... | .................... | .................... | .................... | .................... | |
| E | 5245 hCoV-19/England/BRIS-12FA1B/2020\|EPI_ISL_488419\|20 | .................... | .................... | .................... | .................... | .................... | .................... | |
| E | 5246 hCoV-19/England/BRIS-12F85D/2020\|EPI_ISL_488420\|20 | .................... | .................... | .................... | .................... | .................... | .................... | |
| E | 5247 hCoV-19/England/BRIS-12E95B/2020\|EPI_ISL_488421\|20 | .................... | .................... | .................... | .................... | .................... | .................... | |
| E | 5248 hCoV-19/England/BRIS-12F44A/2020\|EPI_ISL_488422\|20 | .................... | .............A...... | .................... | .................... | .................... | .................... | |
| E | 5249 hCoV-19/England/BRIS-12FB45/2020\|EPI_ISL_488423\|20 | .................... | .................... | .................... | .................... | .................... | .................... | |
| E | 5250 hCoV-19/England/BRIS-12F9D2/2020\|EPI_ISL_488432\|20 | .................... | .................... | .................... | .................... | .................... | .................... | |
| E | 5251 hCoV-19/England/NORT-295A3D/2020\|EPI_ISL_488457\|20 | .................... | .................... | .................... | .................... | .................... | .................... | |
| E | 5252 hCoV-19/England/NORT-282DEF/2020\|EPI_ISL_488458\|20 | .................... | .................... | .................... | .................... | .................... | .................... | |
| E | 5253 hCoV-19/England/NORT-2837A4/2020\|EPI_ISL_488459\|20 | .................... | .................... | .................... | .................... | .................... | .................... | |
| E | 5254 hCoV-19/England/NORT-283F06/2020\|EPI_ISL_488461\|20 | .................... | .................... | .................... | .................... | .................... | .................... | |
| E | 5255 hCoV-19/England/NORT-2958D9/2020\|EPI_ISL_488462\|20 | .................... | .................... | .................... | .................... | .................... | .................... | |
| E | 5256 hCoV-19/England/NORT-282C2E/2020\|EPI_ISL_488463\|20 | .................... | .................... | .................... | .................... | .................... | .................... | |
| E | 5257 hCoV-19/England/NORT-294CFC/2020\|EPI_ISL_488464\|20 | .................... | .................... | .................... | .................... | .................... | .................... | |
| E | 5258 hCoV-19/England/NORT-2955F0/2020\|EPI_ISL_488465\|20 | .................... | .................... | .................... | .................... | .................... | .................... | |
| E | 5259 hCoV-19/England/NORT-284178/2020\|EPI_ISL_488466\|20 | .................... | .................... | .................... | .................... | .................... | .................... | |
| E | 5260 hCoV-19/England/NORT-294F3F/2020\|EPI_ISL_488467\|20 | .................... | .................... | .................... | .................... | .................... | .................... | |
| E | 5261 hCoV-19/England/NORT-295C73/2020\|EPI_ISL_488468\|20 | .................... | .................... | .................... | .................... | .................... | .................... | |
| E | 5262 hCoV-19/England/NORT-2832C1/2020\|EPI_ISL_488469\|20 | .................... | .................... | .................... | .................... | .................... | .................... | |
| E | 5263 hCoV-19/England/NORT-2849E9/2020\|EPI_ISL_488470\|20 | .................... | .................... | .................... | .................... | .................... | .................... | |
| E | 5264 hCoV-19/England/NORT-281FCC/2020\|EPI_ISL_488471\|20 | .................... | .................... | .................... | .................... | .................... | .................... | |
| E | 5265 hCoV-19/England/NORT-294C47/2020\|EPI_ISL_488472\|20 | .................... | .................... | .................... | .................... | .................... | .................... | |
| E | 5266 hCoV-19/England/NORT-2832EF/2020\|EPI_ISL_488473\|20 | .................... | .................... | .................... | .................... | .................... | .................... | |
| E | 5267 hCoV-19/England/NORT-282DB2/2020\|EPI_ISL_488474\|20 | .................... | .................... | .................... | .................... | .................... | .................... | |
| E | 5268 hCoV-19/England/NORT-284934/2020\|EPI_ISL_488475\|20 | .................... | .................... | .................... | .................... | .................... | .................... | |
| E | 5269 hCoV-19/England/NORT-2838ED/2020\|EPI_ISL_488476\|20 | .................... | .................... | .................... | .................... | .................... | .................... | |
| E | 5270 hCoV-19/England/NORT-295824/2020\|EPI_ISL_488477\|20 | .................... | .................... | .................... | .................... | .................... | .................... | |
| E | 5271 hCoV-19/England/NORT-282365/2020\|EPI_ISL_488478\|20 | .................... | .................... | .................... | .................... | .................... | .................... | |
| E | 5272 hCoV-19/England/NORT-28265D/2020\|EPI_ISL_488479\|20 | .................... | .................... | .................... | .................... | .................... | .................... | |
| E | 5273 hCoV-19/England/NORT-294D62/2020\|EPI_ISL_488480\|20 | .................... | .................... | .................... | .................... | .................... | .................... | |
| E | 5274 hCoV-19/England/NORT-282954/2020\|EPI_ISL_488481\|20 | .................... | .................... | .................... | .................... | .................... | .................... | |
| E | 5275 hCoV-19/England/NORT-282787/2020\|EPI_ISL_488482\|20 | .................... | .................... | .................... | .................... | .................... | .................... | |
| E | 5276 hCoV-19/England/NORT-284A6E/2020\|EPI_ISL_488483\|20 | .................... | .................... | .................... | .................... | .................... | .................... | |
| E | 5277 hCoV-19/England/NORT-283E36/2020\|EPI_ISL_488484\|20 | .................... | .................... | .................... | .................... | .................... | .................... | |
| E | 5278 hCoV-19/England/NORT-284275/2020\|EPI_ISL_488485\|20 | .................... | .................... | .................... | .................... | .................... | .................... | |
| E | 5279 hCoV-19/England/NORT-282B21/2020\|EPI_ISL_488486\|20 | .................... | .................... | .................... | .................... | .................... | .................... | |
| E | 5280 hCoV-19/England/NORT-294F8A/2020\|EPI_ISL_488487\|20 | .................... | .................... | .................... | .................... | .................... | .................... | |
| E | 5281 hCoV-19/England/NORT-2954C6/2020\|EPI_ISL_488488\|20 | .................... | .................... | .................... | .................... | .................... | .................... | |
| E | 5282 hCoV-19/England/NORT-295B94/2020\|EPI_ISL_488489\|20 | .................... | ...................A | .................... | .................... | .................... | .................... | 1 |
| E | 5283 hCoV-19/England/NORT-284688/2020\|EPI_ISL_488490\|20 | .................... | .................... | .................... | .................... | .................... | .................... | |
| E | 5284 hCoV-19/England/NORT-283258/2020\|EPI_ISL_488491\|20 | .................... | .................... | .................... | .................... | .................... | .................... | |
| E | 5285 hCoV-19/England/NORT-2952EA/2020\|EPI_ISL_488494\|20 | .................... | .................... | .................... | .................... | .................... | .................... | |
| E | 5286 hCoV-19/England/NORT-2828B1/2020\|EPI_ISL_488495\|20 | .................... | .................... | .................... | .................... | .................... | .................... | |
| E | 5287 hCoV-19/England/NORT-295806/2020\|EPI_ISL_488496\|20 | .................... | .................... | .................... | .................... | .................... | .................... | |
| E | 5288 hCoV-19/England/NORT-294D17/2020\|EPI_ISL_488497\|20 | .................... | .................... | .................... | .................... | .................... | .................... | |
| E | 5289 hCoV-19/England/NORT-28273C/2020\|EPI_ISL_488498\|20 | .................... | .................... | .................... | .................... | .................... | .................... | |
| E | 5290 hCoV-19/Scotland/GCVR-17142B/2020\|EPI_ISL_489590\|2 | .................... | ...................A | .................... | .................... | .................... | .................... | 1 |
| E | 5291 hCoV-19/Scotland/GCVR-171519/2020\|EPI_ISL_489591\|2 | .................... | .................... | .................... | .................... | .................... | .................... | |
| E | 5292 hCoV-19/Scotland/GCVR-1706DB/2020\|EPI_ISL_489593\|2 | .................... | .A.................. | .................... | .................... | .................... | .................... | |
| E | 5293 hCoV-19/Scotland/GCVR-16FEF5/2020\|EPI_ISL_489594\|2 | .................... | .................... | .................... | .................... | .................... | .................... | |
| E | 5294 hCoV-19/Scotland/GCVR-16FC55/2020\|EPI_ISL_489595\|2 | .................... | .................... | .................... | .................... | .................... | .................... | |
| E | 5295 hCoV-19/Scotland/GCVR-171731/2020\|EPI_ISL_489596\|2 | .................... | .................... | .................... | .................... | .................... | .................... | |
| E | 5296 hCoV-19/Scotland/GCVR-16FD25/2020\|EPI_ISL_489597\|2 | .................... | ...................A | .................... | .................... | .................... | .................... | 1 |
| E | 5297 hCoV-19/Scotland/GCVR-16FAD3/2020\|EPI_ISL_489598\|2 | .................... | .................... | .................... | .................... | .................... | .................... | |
| E | 5298 hCoV-19/Scotland/GCVR-171959/2020\|EPI_ISL_489599\|2 | .................... | .................... | .................... | .................... | .................... | .................... | |
| E | 5299 hCoV-19/USA/FL-BPHL-0336/2020\|EPI_ISL_489745\|2020- | .................... | .................... | .................... | .................... | .................... | .................... | |
| E | 5300 hCoV-19/USA/FL-BPHL-0338/2020\|EPI_ISL_489747\|2020- | .................... | .................... | .................... | .................... | .................... | .................... | |
| E | 5301 hCoV-19/USA/FL-BPHL-0343/2020\|EPI_ISL_489752\|2020- | .................... | .................... | .................... | .................... | .................... | .................... | |
| E | 5302 hCoV-19/USA/FL-BPHL-0344/2020\|EPI_ISL_489753\|2020- | .................... | .................... | .................... | .................... | .................... | .................... | |
| E | 5303 hCoV-19/USA/FL-BPHL-0345/2020\|EPI_ISL_489754\|2020- | .................... | .................... | .................... | .................... | ..NN................ | .................... | |
| E | 5304 hCoV-19/USA/FL-BPHL-0379/2020\|EPI_ISL_489788\|2020- | .................... | .................... | .................... | .................... | .................... | ...A................ | |
| E | 5305 hCoV-19/USA/FL-BPHL-0380/2020\|EPI_ISL_489789\|2020- | .................... | .................... | .................... | .................... | .................... | ...A................ | |
| E | 5306 hCoV-19/USA/FL-BPHL-0381/2020\|EPI_ISL_489790\|2020- | .................... | .................... | .................... | .................... | .................... | .................... | |
| E | 5307 hCoV-19/USA/FL-BPHL-0382/2020\|EPI_ISL_489791\|2020- | .................... | .................... | .................... | .................... | .................... | .................... | |
| E | 5308 hCoV-19/USA/FL-BPHL-0383/2020\|EPI_ISL_489792\|2020- | .................... | .................... | .................... | .................... | .................... | .................... | |
| E | 5309 hCoV-19/USA/FL-BPHL-0384/2020\|EPI_ISL_489793\|2020- | .................... | .................... | .................... | .................... | .................... | .................... | |
| E | 5310 hCoV-19/USA/FL-BPHL-0385/2020\|EPI_ISL_489794\|2020- | .................... | .................... | .................... | .................... | .................... | .................... | |
| E | 5311 hCoV-19/USA/FL-BPHL-0386/2020\|EPI_ISL_489795\|2020- | .................... | .................... | .................... | .................... | .................... | .................... | |
| E | 5312 hCoV-19/USA/FL-BPHL-0387/2020\|EPI_ISL_489796\|2020- | .................... | .................... | .................... | .................... | .................... | ...A................ | |
| E | 5313 hCoV-19/USA/FL-BPHL-0388/2020\|EPI_ISL_489797\|2020- | .................... | .................... | .................... | .................... | .................... | ...A................ | |
| E | 5314 hCoV-19/USA/FL-BPHL-0390/2020\|EPI_ISL_489799\|2020- | .................... | .................... | .................... | .................... | .................... | .................... | |
| E | 5315 hCoV-19/England/NOTT-112714/2020\|EPI_ISL_490570\|20 | .................... | .................... | .................... | .................... | .................... | .................... | |
| E | 5316 hCoV-19/England/NOTT-112723/2020\|EPI_ISL_490571\|20 | .................... | .................... | .................... | .................... | .................... | .................... | |
| E | 5317 hCoV-19/England/NOTT-112741/2020\|EPI_ISL_490573\|20 | .................... | .................... | .................... | .................... | .................... | .................... | |

| | | | | | | | | |
|---|---|---|---|---|---|---|---|---|
| E | 5318 | hCoV-19/England/NOTT-112750/2020\|EPI_ISL_490574\|20 | .................... | .................... | ..................... | .................... | .................... | .................... |
| E | 5319 | hCoV-19/England/NOTT-11276F/2020\|EPI_ISL_490575\|20 | .................... | .................... | ..................... | .................... | .................... | .................... |
| E | 5320 | hCoV-19/England/NOTT-11277E/2020\|EPI_ISL_490576\|20 | .................... | .................... | ..................... | .................... | .................... | .................... |
| E | 5321 | hCoV-19/England/NOTT-11278D/2020\|EPI_ISL_490577\|20 | .................... | .................... | ..................... | .................... | .................... | .................... |
| E | 5322 | hCoV-19/England/NOTT-1127AB/2020\|EPI_ISL_490579\|20 | .................... | .................... | ..................... | .................... | .................... | .................... |
| E | 5323 | hCoV-19/England/NOTT-1127BA/2020\|EPI_ISL_490580\|20 | .................... | .................... | ..................... | .................... | .................... | .................... |
| E | 5324 | hCoV-19/England/SHEF-C1742/2020\|EPI_ISL_490581\|202 | .................... | .................... | ..................... | .................... | .................... | .................... |
| E | 5325 | hCoV-19/England/SHEF-C1D04/2020\|EPI_ISL_490585\|202 | .................... | .................... | ..................... | .................... | .................... | .................... |
| E | 5326 | hCoV-19/England/SHEF-C200A/2020\|EPI_ISL_490586\|202 | .................... | .................... | ..................... | .................... | .................... | .................... |
| E | 5327 | hCoV-19/England/SHEF-C2107/2020\|EPI_ISL_490587\|202 | .................... | .................... | ..................... | .................... | .................... | .................... |
| E | 5328 | hCoV-19/England/SHEF-C328B/2020\|EPI_ISL_490588\|202 | .................... | .................... | ..................... | .................... | .................... | .................... |
| E | 5329 | hCoV-19/England/SHEF-C36AD/2020\|EPI_ISL_490589\|202 | .................... | .................... | ..................... | .................... | .................... | .................... |
| E | 5330 | hCoV-19/England/SHEF-C390E/2020\|EPI_ISL_490591\|202 | .................... | .................... | ..................... | .................... | .................... | .................... |
| E | 5331 | hCoV-19/England/SHEF-C5571/2020\|EPI_ISL_490595\|202 | .................... | .................... | ..................... | .................... | .................... | .................... |
| E | 5332 | hCoV-19/England/SHEF-C59A2/2020\|EPI_ISL_490598\|202 | .................... | .................... | ..................... | .................... | .................... | .................... |
| E | 5333 | hCoV-19/Wales/PHWC-16583D/2020\|EPI_ISL_490731\|2020 | .................... | .................... | ..................... | .................... | .................... | .................... |
| E | 5334 | hCoV-19/Wales/PHWC-16590D/2020\|EPI_ISL_490740\|2020 | .................... | .................... | ..................... | .................... | .................... | .................... |
| E | 5335 | hCoV-19/Wales/PHWC-165967/2020\|EPI_ISL_490745\|2020 | .................... | .................... | ..................... | .................... | .................... | .................... |
| E | 5336 | hCoV-19/Wales/PHWC-165994/2020\|EPI_ISL_490747\|2020 | .................... | .................... | ..................... | .................... | .................... | .................... |
| E | 5337 | hCoV-19/Wales/PHWC-165B43/2020\|EPI_ISL_490770\|2020 | .................... | .................... | ..................... | .................... | .................... | .................... |
| E | 5338 | hCoV-19/Wales/PHWC-165B8F/2020\|EPI_ISL_490773\|2020 | .................... | .................... | ..................... | .................... | .................... | .................... |
| E | 5339 | hCoV-19/Wales/PHWC-165C04/2020\|EPI_ISL_490781\|2020 | .................... | .................... | ..................... | .................... | .................... | .................... |
| E | 5340 | hCoV-19/Wales/PHWC-165C13/2020\|EPI_ISL_490782\|2020 | .................... | .................... | ..................... | .................... | .................... | .................... |
| E | 5341 | hCoV-19/Wales/PHWC-165C40/2020\|EPI_ISL_490783\|2020 | .................... | .................... | ..................... | .................... | .................... | .................... |
| E | 5342 | hCoV-19/Wales/PHWC-165C9B/2020\|EPI_ISL_490787\|2020 | .................... | .................... | ..................... | .................... | .................... | .................... |
| E | 5343 | hCoV-19/Wales/PHWC-165CAA/2020\|EPI_ISL_490788\|2020 | .................... | .................... | ..................... | .................... | .................... | .................... |
| E | 5344 | hCoV-19/Wales/PHWC-165CD7/2020\|EPI_ISL_490791\|2020 | .................... | .................... | ..................... | .................... | .................... | .................... |
| E | 5345 | hCoV-19/Wales/PHWC-165D10/2020\|EPI_ISL_490795\|2020 | .................... | .................... | ..................... | .................... | .................... | .................... |
| E | 5346 | hCoV-19/Wales/PHWC-165D2F/2020\|EPI_ISL_490796\|2020 | .................... | .................... | ..................... | .................... | .................... | .................... |
| E | 5347 | hCoV-19/Wales/PHWC-165D98/2020\|EPI_ISL_490799\|2020 | .................... | .................... | ..................... | .................... | .................... | .................... |
| E | 5349 | hCoV-19/Wales/PHWC-166DD3/2020\|EPI_ISL_494115\|2020 | .................... | .................... | ..................... | .................... | .................... | .................... |
| E | 5350 | hCoV-19/Wales/PHWC-166DF1/2020\|EPI_ISL_494117\|2020 | .................... | .................... | ..................... | .................... | .................... | .................... |
| E | 5351 | hCoV-19/Wales/PHWC-166E1C/2020\|EPI_ISL_494118\|2020 | .................... | .................... | ..................... | .................... | .................... | .................... |
| E | 5353 | hCoV-19/Wales/PHWC-166E58/2020\|EPI_ISL_494121\|2020 | .................... | .................... | ..................... | .................... | .................... | .................... |
| E | 5354 | hCoV-19/USA/FL-BPHL-0494/2020\|EPI_ISL_495400\|2020- | .................... | .................... | ..................... | .................... | .................... | .................... |
| E | 5355 | hCoV-19/USA/FL-BPHL-0496/2020\|EPI_ISL_495402\|2020- | .................... | .................... | ..................... | .................... | .................... | .................... |
| E | 5356 | hCoV-19/USA/FL-BPHL-0497/2020\|EPI_ISL_495403\|2020- | .................... | .................... | ..................... | .................... | .................... | .................... |
| E | 5358 | hCoV-19/USA/FL-BPHL-0500/2020\|EPI_ISL_495406\|2020- | .................... | .................... | ..................... | .................... | .................... | .................... |
| E | 5359 | hCoV-19/USA/FL-BPHL-0501/2020\|EPI_ISL_495407\|2020- | .................... | .................... | ..................... | .................... | .................... | .................... |
| E | 5360 | hCoV-19/USA/FL-BPHL-0503/2020\|EPI_ISL_495409\|2020- | .................... | .................... | ..................... | .................... | .................... | .................... |
| E | 5361 | hCoV-19/Turkey/KU-001/2020\|EPI_ISL_495411\|2020-05- | .................... | .................... | ..................... | .................... | .................... | .................... |
| E | 5362 | hCoV-19/Turkey/KU-002/2020\|EPI_ISL_495412\|2020-05- | .................... | .................... | ..................... | .................... | .................... | .................... |
| E | 5363 | hCoV-19/Turkey/KU-003/2020\|EPI_ISL_495413\|2020-05- | .................... | .................... | ..................... | .................... | .................... | .................... |
| E | 5364 | hCoV-19/Turkey/KU-004/2020\|EPI_ISL_495414\|2020-05- | .................... | .................... | ..................... | .................... | .................... | .................... |
| E | 5365 | hCoV-19/Turkey/KU-006/2020\|EPI_ISL_495416\|2020-05- | .................... | .................... | ..................... | .................... | .................... | .................... |
| E | 5366 | hCoV-19/Turkey/KU-007/2020\|EPI_ISL_495417\|2020-05- | .................... | .................... | ..................... | .................... | .................... | .................... |
| E | 5367 | hCoV-19/Turkey/KU-008/2020\|EPI_ISL_495418\|2020-05- | .................... | .................... | ..................... | .................... | .................... | .................... |
| E | 5368 | hCoV-19/Turkey/KU-032/2020\|EPI_ISL_495442\|2020-05- | .................... | .................... | ..................... | .................... | .................... | .................... |
| E | 5369 | hCoV-19/Turkey/KU-033/2020\|EPI_ISL_495443\|2020-04- | .................... | .................... | ..................... | .................... | .................... | .................... |
| E | 5370 | hCoV-19/Turkey/KU-035/2020\|EPI_ISL_495445\|2020-06- | .................... | .................... | ..................... | .................... | .................... | .................... |
| E | 5371 | hCoV-19/Turkey/KU-037/2020\|EPI_ISL_495447\|2020-05- | .................... | .................... | ..................... | .................... | .................... | .................... |
| E | 5372 | hCoV-19/Turkey/KU-038/2020\|EPI_ISL_495448\|2020-04- | .................... | .................... | ..................... | .................... | .................... | .................... |
| E | 5373 | hCoV-19/Turkey/KU-039/2020\|EPI_ISL_495449\|2020-05- | .................... | .................... | ..................... | .................... | .................... | .................... |
| E | 5374 | hCoV-19/Turkey/KU-040/2020\|EPI_ISL_495450\|2020-05- | .................... | .................... | ..................... | .................... | .................... | .................... |
| E | 5375 | hCoV-19/Turkey/KU-041/2020\|EPI_ISL_495451\|2020-05- | .................... | .A.................. | ..................... | .................... | .................... | .................... |
| E | 5376 | hCoV-19/Turkey/KU-042/2020\|EPI_ISL_495452\|2020-05- | .................... | .................... | ..................... | .................... | .................... | .................... |
| E | 5377 | hCoV-19/USA/OR-PROV-001/2020\|EPI_ISL_482292\|2020-0 | .................... | .................... | ..................... | .................... | .................... | .................... |
| E | 5378 | hCoV-19/USA/OR-PROV-002/2020\|EPI_ISL_482293\|2020-0 | .................... | .................... | ..................... | .................... | .................... | .................... |
| E | 5379 | hCoV-19/USA/OR-PROV-003/2020\|EPI_ISL_482294\|2020-0 | .................... | .................... | ..................... | .................... | .................... | .................... |
| E | 5380 | hCoV-19/USA/OR-PROV-004/2020\|EPI_ISL_482295\|2020-0 | .................... | .................... | ..................... | .................... | .................... | .................... |
| E | 5381 | hCoV-19/USA/OR-PROV-007/2020\|EPI_ISL_482296\|2020-0 | .................... | .................... | ..................... | .................... | .................... | .................... |
| E | 5382 | hCoV-19/USA/OR-PROV-009/2020\|EPI_ISL_482297\|2020-0 | .................... | .................... | ..................... | .................... | .................... | .................... |
| E | 5383 | hCoV-19/USA/OR-PROV-011/2020\|EPI_ISL_482298\|2020-0 | .................... | .................... | ..................... | .................... | .................... | .................... |
| E | 5384 | hCoV-19/USA/OR-PROV-012/2020\|EPI_ISL_482299\|2020-0 | .................... | .................... | ..................... | .................... | .................... | .................... |
| E | 5385 | hCoV-19/USA/CA-ALSR-1093-SAN/2020\|EPI_ISL_483520\|2 | .................... | .A.................. | ..................... | .................... | .................... | .................... |
| E | 5386 | hCoV-19/USA/CA-ALSR-1094-SAN/2020\|EPI_ISL_483521\|2 | .................... | .A.................. | ..................... | .................... | .................... | .................... |
| E | 5387 | hCoV-19/USA/CA-ALSR-1098-SAN/2020\|EPI_ISL_483525\|2 | .................... | .................... | ..................... | .................... | .................... | .................... |
| E | 5388 | hCoV-19/Bahrein/BAH-21/2020\|EPI_ISL_483562\|2020-04 | .................... | .................... | ..................... | .................... | .................... | .................... |
| E | 5389 | hCoV-19/Bahrein/BAH-22/2020\|EPI_ISL_483563\|2020-04 | .................... | .................... | ..................... | .................... | .................... | .................... |
| E | 5390 | hCoV-19/Bahrein/BAH-23/2020\|EPI_ISL_483564\|2020-04 | .................... | .................... | ..................... | .................... | .................... | .................... |
| E | 5391 | hCoV-19/Bahrein/BAH-27/2020\|EPI_ISL_483565\|2020-04 | .................... | .................... | ..................... | .................... | .................... | .................... |
| E | 5392 | hCoV-19/Spain/PV-HUB-56597693/2020\|EPI_ISL_483566\| | .................... | .................... | ..................... | .................... | .................... | .................... |
| E | 5393 | hCoV-19/Spain/PV-HUB-50399463/2020\|EPI_ISL_483570\| | .................... | .................... | ..................... | .................... | .................... | .................... |
| E | 5394 | hCoV-19/Spain/PV-HUB-51235493/2020\|EPI_ISL_483571\| | .................... | .................... | ..................... | .................... | .................... | .................... |
| E | 5395 | hCoV-19/Spain/PV-HUB-56574605/2020\|EPI_ISL_483572\| | .................... | .................... | ..................... | .................... | .................... | .................... |
| E | 5396 | hCoV-19/Spain/PV-HUB-52580776/2020\|EPI_ISL_483573\| | .................... | .................... | ..................... | .................... | .................... | .................... |
| E | 5397 | hCoV-19/Singapore/557/2020\|EPI_ISL_483580\|2020-05- | .................... | .................... | ..................... | .................... | .................... | .................... |
| E | 5398 | hCoV-19/Singapore/558/2020\|EPI_ISL_483581\|2020-04- | .................... | .................... | ..................... | .................... | .................... | .................... |
| E | 5399 | hCoV-19/Singapore/559/2020\|EPI_ISL_483582\|2020-06- | .................... | .................... | ..................... | .................... | .................... | .................... |
| E | 5400 | hCoV-19/Singapore/560/2020\|EPI_ISL_483583\|2020-04- | .................... | .................... | ..................... | .................... | .................... | .................... |

| | | | | | | | | |
|---|---|---|---|---|---|---|---|---|
| E | 5401 | hCoV-19/Singapore/561/2020\|EPI_ISL_483584\|2020-05- | ···················· | ···················· | ····················· | ···················· | ···················· | ···················· |
| E | 5402 | hCoV-19/Singapore/562/2020\|EPI_ISL_483585\|2020-04- | ···················· | ···················· | ····················· | ···················· | ···················· | ···················· |
| E | 5403 | hCoV-19/Singapore/563/2020\|EPI_ISL_483586\|2020-05- | ···················· | ···················· | ····················· | ···················· | ···················· | ···················· |
| E | 5404 | hCoV-19/Singapore/564/2020\|EPI_ISL_483587\|2020-04- | ···················· | ···················· | ····················· | ···················· | ···················· | ···················· |
| E | 5405 | hCoV-19/Singapore/565/2020\|EPI_ISL_483588\|2020-05- | ···················· | ···················· | ····················· | ···················· | ···················· | ···················· |
| E | 5406 | hCoV-19/Singapore/566/2020\|EPI_ISL_483589\|2020-04- | ···················· | ···················· | ····················· | ···················· | ···················· | ···················· |
| E | 5407 | hCoV-19/Singapore/567/2020\|EPI_ISL_483590\|2020-06- | ···················· | ···················· | ····················· | ···················· | ···················· | ···················· |
| E | 5408 | hCoV-19/Singapore/568/2020\|EPI_ISL_483591\|2020-05- | ···················· | ···················· | ····················· | ···················· | ···················· | ···················· |
| E | 5409 | hCoV-19/Singapore/569/2020\|EPI_ISL_483592\|2020-05- | ···················· | ···················· | ····················· | ···················· | ···················· | ···················· |
| E | 5410 | hCoV-19/Singapore/573/2020\|EPI_ISL_483596\|2020-04- | ···················· | ···················· | ····················· | ···················· | ···················· | ···················· |
| E | 5411 | hCoV-19/Singapore/574/2020\|EPI_ISL_483597\|2020-04- | ···················· | ···················· | ····················· | ···················· | ···················· | ···················· |
| E | 5412 | hCoV-19/Singapore/575/2020\|EPI_ISL_483598\|2020-05- | ···················· | ···················· | ····················· | ···················· | ···················· | ···················· |
| E | 5413 | hCoV-19/Singapore/576/2020\|EPI_ISL_483599\|2020-05- | ···················· | ···················· | ····················· | ···················· | ···················· | ···················· |
| E | 5414 | hCoV-19/USA/WI-UW-475/2020\|EPI_ISL_484850\|2020-06- | ···················· | ···················· | ····················· | ···················· | ···················· | ···················· |
| E | 5415 | hCoV-19/USA/IL-UW-478/2020\|EPI_ISL_484853\|2020-06- | ···················· | ···················· | ····················· | ···················· | ···················· | ···················· |
| E | 5416 | hCoV-19/USA/WI-UW-479/2020\|EPI_ISL_484854\|2020-06- | ···················· | ···················· | ····················· | ···················· | ···················· | ···················· |
| E | 5417 | hCoV-19/USA/WI-UW-480/2020\|EPI_ISL_484855\|2020-06- | ···················· | ···················· | ····················· | ···················· | ···················· | ···················· |
| E | 5418 | hCoV-19/USA/WI-UW-481/2020\|EPI_ISL_484856\|2020-06- | ···················· | ···················· | ····················· | ···················· | ···················· | ···················· |
| E | 5419 | hCoV-19/USA/WI-UW-518/2020\|EPI_ISL_484893\|2020-06- | ···················· | ···················· | ····················· | ···················· | ···················· | ···················· |
| E | 5420 | hCoV-19/USA/WI-UW-519/2020\|EPI_ISL_484894\|2020-06- | ···················· | ···················· | ····················· | ···················· | ···················· | ···················· |
| E | 5421 | hCoV-19/USA/WI-UW-520/2020\|EPI_ISL_484895\|2020-06- | ···················· | ···················· | ····················· | ···················· | ···················· | ···················· |
| E | 5422 | hCoV-19/USA/WI-UW-521/2020\|EPI_ISL_484896\|2020-06- | ···················· | ···················· | ····················· | ···················· | ···················· | ···················· |
| E | 5423 | hCoV-19/USA/WI-UW-522/2020\|EPI_ISL_484897\|2020-06- | ···················· | ···················· | ····················· | ···················· | ···················· | ···················· |
| E | 5424 | hCoV-19/USA/WI-UW-523/2020\|EPI_ISL_484898\|2020-06- | ···················· | ···················· | ····················· | ···················· | ···················· | ···················· |
| E | 5425 | hCoV-19/USA/WI-UW-524/2020\|EPI_ISL_484899\|2020-06- | ···················· | ···················· | ····················· | ···················· | ···················· | ···················· |
| E | 5426 | hCoV-19/Nigeria/OY260-CV55/2020\|EPI_ISL_487101\|202 | ···················· | ···················· | ····················· | ···················· | ···················· | ···················· |
| E | 5427 | hCoV-19/Nigeria/EK220-CV54/2020\|EPI_ISL_487102\|202 | ···················· | ···················· | ····················· | ···················· | ···················· | ···················· |
| E | 5428 | hCoV-19/Nigeria/KW326-CV52/2020\|EPI_ISL_487103\|202 | ···················· | ···················· | ····················· | ···················· | ···················· | ···················· |
| E | 5429 | hCoV-19/Nigeria/EK084-CV50/2020\|EPI_ISL_487105\|202 | ···················· | ···················· | ····················· | ···················· | ···················· | ···················· |
| E | 5430 | hCoV-19/Nigeria/ON191-CV49/2020\|EPI_ISL_487106\|202 | ···················· | ······A············· | ····················· | ···················· | ···················· | ···················· |
| E | 5431 | hCoV-19/Nigeria/KW298-CV48/2020\|EPI_ISL_487107\|202 | ···················· | ···················· | ····················· | ···················· | ···················· | ···················· |
| E | 5432 | hCoV-19/Nigeria/EK082-CV47/2020\|EPI_ISL_487108\|202 | ···················· | ···················· | ····················· | ···················· | ···················· | ···················· |
| E | 5433 | hCoV-19/Nigeria/OS589-CV46/2020\|EPI_ISL_487109\|202 | ···················· | ···················· | ····················· | ···················· | ···················· | ···················· |
| E | 5434 | hCoV-19/Nigeria/OS524-CV45/2020\|EPI_ISL_487110\|202 | ···················· | ···················· | ····················· | ···················· | ···················· | ···················· |
| E | 5435 | hCoV-19/Nigeria/ON505-CV43/2020\|EPI_ISL_487112\|202 | ···················· | ···················· | ····················· | ···················· | ···················· | ···················· |
| E | 5436 | hCoV-19/Nigeria/OS294-CV42/2020\|EPI_ISL_487113\|202 | ···················· | ···················· | ····················· | ···················· | ···················· | ···················· |
| E | 5437 | hCoV-19/England/BRIS-12F32F/2020\|EPI_ISL_488400\|20 | ···················· | ···················· | ····················· | ···················· | ···················· | ···················· |
| E | 5438 | hCoV-19/England/BRIS-12E5FD/2020\|EPI_ISL_488401\|20 | ···················· | ···················· | ····················· | ···················· | ···················· | ···················· |
| E | 5439 | hCoV-19/England/BRIS-12F064/2020\|EPI_ISL_488402\|20 | ···················· | ···················· | ····················· | ···················· | ···················· | ···················· |
| E | 5440 | hCoV-19/England/BRIS-12ED6E/2020\|EPI_ISL_488403\|20 | ···················· | ···················· | ····················· | ···················· | ···················· | ···················· |
| E | 5441 | hCoV-19/England/BRIS-12F969/2020\|EPI_ISL_488404\|20 | ···················· | ···················· | ····················· | ···················· | ···················· | ···················· |
| E | 5442 | hCoV-19/England/BRIS-12EE7A/2020\|EPI_ISL_488405\|20 | ···················· | ···················· | ····················· | ···················· | ···················· | ···················· |
| E | 5443 | hCoV-19/England/BRIS-12EBA0/2020\|EPI_ISL_488406\|20 | ···················· | ···················· | ····················· | ···················· | ···················· | ···················· |
| E | 5444 | hCoV-19/England/BRIS-12E7AC/2020\|EPI_ISL_488407\|20 | ···················· | ···················· | ····················· | ···················· | ···················· | ···················· |
| E | 5445 | hCoV-19/England/BRIS-12F95A/2020\|EPI_ISL_488410\|20 | ···················· | ···················· | ····················· | ···················· | ···················· | ···················· |
| E | 5446 | hCoV-19/England/BRIS-12E4B4/2020\|EPI_ISL_488412\|20 | ···················· | ···················· | ····················· | ···················· | ···················· | ···················· |
| E | 5447 | hCoV-19/England/BRIS-12F529/2020\|EPI_ISL_488425\|20 | ···················· | ···················· | ····················· | ···················· | ···················· | ···················· |
| E | 5448 | hCoV-19/England/BRIS-12EC9E/2020\|EPI_ISL_488426\|20 | ···················· | ···················· | ····················· | ···················· | ···················· | ···················· |
| E | 5449 | hCoV-19/England/BRIS-12F18F/2020\|EPI_ISL_488427\|20 | ···················· | ···················· | ····················· | ···················· | ···················· | ···················· |
| E | 5450 | hCoV-19/England/BRIS-12E94C/2020\|EPI_ISL_488429\|20 | ···················· | ···················· | ····················· | ···················· | ···················· | ···················· |
| E | 5451 | hCoV-19/England/BRIS-12E9F1/2020\|EPI_ISL_488430\|20 | ···················· | ···················· | ····················· | ···················· | ···················· | ···················· |
| E | 5452 | hCoV-19/England/BRIS-12EAEF/2020\|EPI_ISL_488431\|20 | ···················· | ···················· | ····················· | ···················· | ···················· | ···················· |
| E | 5453 | hCoV-19/England/BRIS-12E715/2020\|EPI_ISL_488433\|20 | ···················· | ···················· | ····················· | ···················· | ···················· | ···················· |
| E | 5454 | hCoV-19/England/BRIS-12F77E/2020\|EPI_ISL_488434\|20 | ···················· | ···················· | ····················· | ···················· | ···················· | ···················· |
| E | 5455 | hCoV-19/England/BRIS-12F27D/2020\|EPI_ISL_488435\|20 | ···················· | ···················· | ····················· | ···················· | ···················· | ···················· |
| E | 5456 | hCoV-19/England/BRIS-12E706/2020\|EPI_ISL_488436\|20 | ···················· | ···················· | ····················· | ···················· | ···················· | ···················· |
| E | 5457 | hCoV-19/England/BRIS-12EF0E/2020\|EPI_ISL_488437\|20 | ···················· | ···················· | ····················· | ···················· | ···················· | ···················· |
| E | 5458 | hCoV-19/England/BRIS-12EEC5/2020\|EPI_ISL_488438\|20 | ···················· | ···················· | ····················· | ···················· | ···················· | ···················· |
| E | 5459 | hCoV-19/England/BRIS-12FA66/2020\|EPI_ISL_488439\|20 | ···················· | ···················· | ····················· | ···················· | ···················· | ···················· |
| E | 5460 | hCoV-19/England/BRIS-12EFFF/2020\|EPI_ISL_488440\|20 | ···················· | ···················· | ····················· | ···················· | ···················· | ···················· |
| E | 5461 | hCoV-19/England/BRIS-12EA94/2020\|EPI_ISL_488442\|20 | ···················· | ···················· | ····················· | ···················· | ···················· | ···················· |
| E | 5462 | hCoV-19/England/BRIS-12E593/2020\|EPI_ISL_488443\|20 | ···················· | ···················· | ····················· | ···················· | ···················· | ···················· |
| E | 5463 | hCoV-19/England/BRIS-12F653/2020\|EPI_ISL_488444\|20 | ···················· | ···················· | ····················· | ···················· | ···················· | ···················· |
| E | 5464 | hCoV-19/England/BRIS-12F389/2020\|EPI_ISL_488445\|20 | ···················· | ···················· | ····················· | ···················· | ···················· | ···················· |
| E | 5465 | hCoV-19/England/BRIS-12F3B6/2020\|EPI_ISL_488446\|20 | ···················· | ···················· | ····················· | ···················· | ···················· | ···················· |
| E | 5466 | hCoV-19/England/BRIS-12F78D/2020\|EPI_ISL_488447\|20 | ···················· | ···················· | ····················· | ···················· | ···················· | ···················· |
| E | 5467 | hCoV-19/England/BRIS-12E751/2020\|EPI_ISL_488448\|20 | ···················· | ···················· | ····················· | ···················· | ···················· | ···················· |
| E | 5468 | hCoV-19/England/BRIS-12E742/2020\|EPI_ISL_488449\|20 | ···················· | ···················· | ····················· | ···················· | ···················· | ···················· |
| E | 5469 | hCoV-19/England/BRIS-12F5DE/2020\|EPI_ISL_488450\|20 | ···················· | ···················· | ····················· | ···················· | ···················· | ···················· |
| E | 5470 | hCoV-19/England/BRIS-12F3F2/2020\|EPI_ISL_488451\|20 | ···················· | ···················· | ····················· | ···················· | ···················· | ···················· |
| E | 5471 | hCoV-19/England/BRIS-12F073/2020\|EPI_ISL_488452\|20 | ···················· | ···················· | ····················· | ···················· | ···················· | ···················· |
| E | 5472 | hCoV-19/England/BRIS-12F37A/2020\|EPI_ISL_488453\|20 | ···················· | ···················· | ····················· | ···················· | ···················· | ···················· |
| E | 5473 | hCoV-19/England/BRIS-12F9A5/2020\|EPI_ISL_488455\|20 | ···················· | ···················· | ····················· | ···················· | ···················· | ···················· |
| E | 5474 | hCoV-19/England/NORT-283D0C/2020\|EPI_ISL_488456\|20 | ···················· | ···················· | ····················· | ···················· | ···················· | ···················· |
| E | 5475 | hCoV-19/Scotland/GCVR-1718F2/2020\|EPI_ISL_489701\|2 | ···················· | ···················· | ····················· | ···················· | ···················· | ···················· |
| E | 5476 | hCoV-19/Scotland/GCVR-171740/2020\|EPI_ISL_489703\|2 | ···················· | ···················· | ····················· | ···················· | ···················· | ···················· |
| E | 5477 | hCoV-19/Scotland/GCVR-170EC4/2020\|EPI_ISL_489704\|2 | ···················· | ···················· | ····················· | ···················· | ···················· | ···················· |
| E | 5478 | hCoV-19/Scotland/GCVR-17179B/2020\|EPI_ISL_489705\|2 | ···················· | ···················· | ····················· | ···················· | ···················· | ···················· |
| E | 5479 | hCoV-19/Scotland/GCVR-170F2B/2020\|EPI_ISL_489706\|2 | ···················· | ···················· | ····················· | ···················· | ···················· | ···················· |
| E | 5480 | hCoV-19/Czech Republic/NRL_5556/2020\|EPI_ISL_48970 | ···················· | ···················· | ····················· | ···················· | ···················· | ···················· |

| | | | | | | | | | |
|---|---|---|---|---|---|---|---|---|---|
| E | 5481 | hCoV-19/Czech Republic/NRL_5513/2020\|EPI_ISL_48970 | .................... | .................... | .................... | .................... | .................... | .................... | |
| E | 5482 | hCoV-19/USA/FL-BPHL-0301/2020\|EPI_ISL_489710\|2020- | .................... | .................... | .................... | .................... | .................... | .................... | |
| E | 5483 | hCoV-19/USA/FL-BPHL-0302/2020\|EPI_ISL_489711\|2020- | .................... | .................... | .................... | .................... | .................... | .................... | |
| E | 5484 | hCoV-19/USA/FL-BPHL-0303/2020\|EPI_ISL_489712\|2020- | .................... | .................... | .................... | .................... | .................... | .................... | |
| E | 5485 | hCoV-19/USA/FL-BPHL-0307/2020\|EPI_ISL_489716\|2020- | .................... | .................... | .................... | .................... | .................... | .................... | |
| E | 5486 | hCoV-19/USA/FL-BPHL-0308/2020\|EPI_ISL_489717\|2020- | .................... | .................... | .................... | .................... | .................... | .................... | |
| E | 5487 | hCoV-19/USA/FL-BPHL-0309/2020\|EPI_ISL_489718\|2020- | .................... | .................... | .................... | .................... | .................... | .................... | |
| E | 5488 | hCoV-19/USA/FL-BPHL-0311/2020\|EPI_ISL_489720\|2020- | .................... | .................... | .................... | .................... | .................... | .................... | |
| E | 5489 | hCoV-19/USA/FL-BPHL-0313/2020\|EPI_ISL_489722\|2020- | .................... | .................... | .................... | .................... | .................... | .................... | |
| E | 5490 | hCoV-19/USA/FL-BPHL-0314/2020\|EPI_ISL_489723\|2020- | .................... | .................... | .................... | .................... | .................... | .................... | |
| E | 5491 | hCoV-19/USA/FL-BPHL-0315/2020\|EPI_ISL_489724\|2020- | .................... | .................... | .................... | .................... | .................... | .................... | |
| E | 5492 | hCoV-19/USA/FL-BPHL-0318/2020\|EPI_ISL_489727\|2020- | .................... | .................... | .................... | .................... | .................... | .................... | |
| E | 5493 | hCoV-19/USA/FL-BPHL-0319/2020\|EPI_ISL_489728\|2020- | .................... | .................... | .................... | .................... | .................... | .................... | |
| E | 5494 | hCoV-19/USA/FL-BPHL-0320/2020\|EPI_ISL_489729\|2020- | .................... | .................... | .................... | .................... | .................... | .................... | |
| E | 5495 | hCoV-19/USA/FL-BPHL-0321/2020\|EPI_ISL_489730\|2020- | .................... | .................... | .................... | .................... | .................... | .................... | |
| E | 5496 | hCoV-19/USA/FL-BPHL-0322/2020\|EPI_ISL_489731\|2020- | .................... | .................... | .................... | .................... | .................... | .................... | |
| E | 5497 | hCoV-19/USA/FL-BPHL-0324/2020\|EPI_ISL_489733\|2020- | .................... | .................... | .................... | .................... | .................... | .................... | |
| E | 5498 | hCoV-19/USA/FL-BPHL-0325/2020\|EPI_ISL_489734\|2020- | .................... | .................... | .................... | .................... | .................... | .................... | |
| E | 5499 | hCoV-19/USA/FL-BPHL-0328/2020\|EPI_ISL_489737\|2020- | .................... | .................... | .................... | .................... | .................... | .................... | |
| E | 5500 | hCoV-19/USA/FL-BPHL-0330/2020\|EPI_ISL_489739\|2020- | .................... | .................... | .................... | .................... | .................... | .................... | |
| E | 5502 | hCoV-19/USA/FL-BPHL-0347/2020\|EPI_ISL_489756\|2020- | .................... | .................... | .................... | .................... | .................... | .................... | |
| E | 5503 | hCoV-19/USA/FL-BPHL-0349/2020\|EPI_ISL_489758\|2020- | .................... | .................... | .................... | .................... | .................... | .................... | |
| E | 5504 | hCoV-19/USA/FL-BPHL-0350/2020\|EPI_ISL_489759\|2020- | .................... | .................... | .................... | .................... | .................... | .................... | |
| E | 5505 | hCoV-19/USA/FL-BPHL-0351/2020\|EPI_ISL_489760\|2020- | .................... | .................... | .................... | .................... | .................... | .................... | |
| E | 5506 | hCoV-19/USA/FL-BPHL-0352/2020\|EPI_ISL_489761\|2020- | .................... | .................... | .................... | .................... | .................... | .................... | |
| E | 5507 | hCoV-19/USA/FL-BPHL-0353/2020\|EPI_ISL_489762\|2020- | .................... | .................... | .................... | .................... | .................... | .................... | |
| E | 5508 | hCoV-19/USA/FL-BPHL-0354/2020\|EPI_ISL_489763\|2020- | .................... | .................... | .................... | .................... | .................... | .................... | |
| E | 5509 | hCoV-19/USA/FL-BPHL-0355/2020\|EPI_ISL_489764\|2020- | .................... | ............A....... | .................... | .................... | .................... | .................... | |
| E | 5510 | hCoV-19/USA/FL-BPHL-0356/2020\|EPI_ISL_489765\|2020- | .................... | .................... | .................... | .................... | .................... | .................... | |
| E | 5511 | hCoV-19/USA/FL-BPHL-0358/2020\|EPI_ISL_489767\|2020- | .................... | .................... | .................... | .................... | .................... | .................... | |
| E | 5512 | hCoV-19/USA/FL-BPHL-0360/2020\|EPI_ISL_489769\|2020- | .................... | .................... | .................... | .................... | .................... | .................... | |
| E | 5513 | hCoV-19/USA/FL-BPHL-0361/2020\|EPI_ISL_489770\|2020- | .................... | .................... | .................... | .................... | .................... | .................... | |
| E | 5514 | hCoV-19/USA/FL-BPHL-0363/2020\|EPI_ISL_489772\|2020- | .................... | .................... | .................... | .................... | .................... | .................... | |
| E | 5515 | hCoV-19/USA/FL-BPHL-0364/2020\|EPI_ISL_489773\|2020- | .................... | .................... | .................... | .................... | .................... | .................... | |
| E | 5516 | hCoV-19/USA/FL-BPHL-0368/2020\|EPI_ISL_489777\|2020- | .................... | .................... | .................... | .................... | NNN................. | .................... | 1 |
| E | 5517 | hCoV-19/USA/FL-BPHL-0369/2020\|EPI_ISL_489778\|2020- | .................... | .................... | .................... | .................... | .................... | .................... | |
| E | 5518 | hCoV-19/USA/FL-BPHL-0371/2020\|EPI_ISL_489780\|2020- | .................... | .................... | .................... | .................... | .................... | .................... | |
| E | 5519 | hCoV-19/USA/FL-BPHL-0372/2020\|EPI_ISL_489781\|2020- | .................... | .................... | .................... | .................... | .................... | .................... | |
| E | 5520 | hCoV-19/USA/FL-BPHL-0373/2020\|EPI_ISL_489782\|2020- | .................... | .................... | .................... | .................... | .................... | .................... | |
| E | 5521 | hCoV-19/USA/FL-BPHL-0375/2020\|EPI_ISL_489784\|2020- | .................... | .................... | .................... | .................... | .................... | .................... | |
| E | 5522 | hCoV-19/USA/FL-BPHL-0376/2020\|EPI_ISL_489785\|2020- | .................... | .................... | .................... | .................... | .................... | .................... | |
| E | 5523 | hCoV-19/USA/FL-BPHL-0377/2020\|EPI_ISL_489786\|2020- | .................... | .................... | .................... | .................... | .................... | .................... | |
| E | 5524 | hCoV-19/USA/FL-BPHL-0378/2020\|EPI_ISL_489787\|2020- | .................... | .................... | .................... | .................... | .................... | ..A................. | |
| E | 5525 | hCoV-19/Scotland/CVR3935/2020\|EPI_ISL_490700\|2020- | .................... | .................... | .................... | .................... | .................... | .................... | |
| E | 5526 | hCoV-19/Scotland/CVR3936/2020\|EPI_ISL_490701\|2020- | .................... | .................... | .................... | .................... | .................... | .................... | |
| E | 5527 | hCoV-19/Scotland/CVR3938/2020\|EPI_ISL_490702\|2020- | .................... | .................... | .................... | .................... | .................... | .................... | |
| E | 5528 | hCoV-19/Scotland/CVR3939/2020\|EPI_ISL_490703\|2020- | .................... | .................... | .................... | .................... | .................... | .................... | |
| E | 5529 | hCoV-19/Scotland/CVR3940/2020\|EPI_ISL_490704\|2020- | .................... | .................... | .................... | .................... | .................... | .................... | |
| E | 5530 | hCoV-19/Scotland/CVR3941/2020\|EPI_ISL_490705\|2020- | .................... | .................... | .................... | .................... | .................... | .................... | |
| E | 5531 | hCoV-19/Scotland/CVR3942/2020\|EPI_ISL_490706\|2020- | .................... | .................... | .................... | .................... | .................... | .................... | |
| E | 5532 | hCoV-19/Scotland/CVR3943/2020\|EPI_ISL_490707\|2020- | .................... | .................... | .................... | .................... | .................... | .................... | |
| E | 5533 | hCoV-19/Scotland/CVR3945/2020\|EPI_ISL_490709\|2020- | .................... | .................... | .................... | .................... | .................... | .................... | |
| E | 5534 | hCoV-19/Wales/PHWC-1656AC/2020\|EPI_ISL_490712\|2020 | .................... | .................... | .................... | .................... | .................... | .................... | |
| E | 5535 | hCoV-19/Wales/PHWC-165730/2020\|EPI_ISL_490718\|2020 | .................... | .................... | .................... | .................... | .................... | .................... | |
| E | 5536 | hCoV-19/Wales/PHWC-16577C/2020\|EPI_ISL_490722\|2020 | .................... | .................... | .................... | .................... | .................... | .................... | |
| E | 5537 | hCoV-19/Wales/PHWC-16578B/2020\|EPI_ISL_490723\|2020 | .................... | .................... | .................... | .................... | ..N................. | .................... | |
| E | 5538 | hCoV-19/Wales/PHWC-1657E5/2020\|EPI_ISL_490726\|2020 | .................... | .................... | .................... | .................... | .................... | .................... | |
| E | 5539 | hCoV-19/Wales/PHWC-1657F4/2020\|EPI_ISL_490727\|2020 | .................... | .................... | .................... | .................... | .................... | .................... | |
| E | 5540 | hCoV-19/Wales/PHWC-165800/2020\|EPI_ISL_490728\|2020 | .................... | .................... | .................... | .................... | .................... | .................... | |
| E | 5541 | hCoV-19/Wales/PHWC-165879/2020\|EPI_ISL_490733\|2020 | .................... | .................... | .................... | .................... | .................... | .................... | |
| E | 5542 | hCoV-19/Wales/PHWC-165897/2020\|EPI_ISL_490735\|2020 | .................... | .................... | .................... | .................... | .................... | .................... | |
| E | 5543 | hCoV-19/Wales/PHWC-1658F1/2020\|EPI_ISL_490739\|2020 | .................... | .................... | .................... | .................... | .NN................. | .................... | |
| E | 5544 | hCoV-19/Wales/PHWC-1659EF/2020\|EPI_ISL_490751\|2020 | .................... | .................... | .................... | .................... | .................... | .................... | |
| E | 5545 | hCoV-19/Wales/PHWC-165A19/2020\|EPI_ISL_490753\|2020 | .................... | .................... | .................... | .................... | .................... | .................... | |
| E | 5546 | hCoV-19/Wales/PHWC-165A37/2020\|EPI_ISL_490755\|2020 | .................... | .................... | .................... | .................... | .................... | .................... | |
| E | 5547 | hCoV-19/Wales/PHWC-165A73/2020\|EPI_ISL_490759\|2020 | .................... | .................... | .................... | .................... | .................... | .................... | |
| E | 5548 | hCoV-19/Wales/PHWC-165A82/2020\|EPI_ISL_490760\|2020 | .................... | .................... | .................... | .................... | .................... | .................... | |
| E | 5549 | hCoV-19/Wales/PHWC-165AA0/2020\|EPI_ISL_490762\|2020 | .................... | .................... | .................... | .................... | .................... | .................... | |
| E | 5550 | hCoV-19/Wales/PHWC-165ACE/2020\|EPI_ISL_490763\|2020 | .................... | .................... | .................... | .................... | .................... | .................... | |
| E | 5551 | hCoV-19/Wales/PHWC-165B07/2020\|EPI_ISL_490767\|2020 | .................... | .................... | .................... | .................... | .................... | .................... | |
| E | 5552 | hCoV-19/Wales/PHWC-165B34/2020\|EPI_ISL_490769\|2020 | .................... | .................... | .................... | .................... | .................... | .................... | |
| E | 5553 | hCoV-19/Wales/PHWC-1681D5/2020\|EPI_ISL_494211\|2020 | .................... | .................... | .................... | .................... | .................... | .................... | |
| E | 5554 | hCoV-19/Wales/PHWC-1681F3/2020\|EPI_ISL_494212\|2020 | .................... | .................... | .................... | .................... | .................... | .................... | |
| E | 5555 | hCoV-19/Wales/PHWC-16825A/2020\|EPI_ISL_494216\|2020 | .................... | ...A................ | .................... | .................... | .................... | .................... | |
| E | 5556 | hCoV-19/Wales/PHWC-168269/2020\|EPI_ISL_494217\|2020 | .................... | .................... | .................... | .................... | .................... | .................... | |
| E | 5557 | hCoV-19/Wales/PHWC-168278/2020\|EPI_ISL_494218\|2020 | .................... | .................... | .................... | .................... | .................... | .................... | |
| E | 5558 | hCoV-19/Wales/PHWC-1682A5/2020\|EPI_ISL_494221\|2020 | .................... | .................... | .................... | .................... | .................... | .................... | |
| E | 5559 | hCoV-19/Wales/PHWC-1682E1/2020\|EPI_ISL_494224\|2020 | .................... | .................... | .................... | .................... | .................... | .................... | |
| E | 5560 | hCoV-19/Wales/PHWC-16830C/2020\|EPI_ISL_494225\|2020 | .................... | .................... | .................... | .................... | .................... | .................... | |
| E | 5561 | hCoV-19/Wales/PHWC-16831B/2020\|EPI_ISL_494226\|2020 | .................... | .................... | .................... | .................... | .................... | .................... | |

| | | | | | | | | |
|---|---|---|---|---|---|---|---|---|
| E | 5562 | hCoV-19/Wales/PHWC-16832A/2020\|EPI_ISL_494227\|2020 | ···················· | ···················· | ···················· | ···················· | ···················· | ···················· |
| E | 5563 | hCoV-19/Wales/PHWC-168339/2020\|EPI_ISL_494228\|2020 | ···················· | ···················· | ···················· | ···················· | ···················· | ···················· |
| E | 5564 | hCoV-19/Wales/PHWC-168348/2020\|EPI_ISL_494229\|2020 | ···················· | ···················· | ···················· | ···················· | ···················· | ···················· |
| E | 5565 | hCoV-19/Wales/PHWC-168357/2020\|EPI_ISL_494230\|2020 | ···················· | ···················· | ···················· | ···················· | ···················· | ···················· |
| E | 5566 | hCoV-19/Wales/PHWC-168366/2020\|EPI_ISL_494231\|2020 | ···················· | ···················· | ···················· | ···················· | ···················· | ···················· |
| E | 5567 | hCoV-19/Wales/PHWC-168375/2020\|EPI_ISL_494232\|2020 | ···················· | ···················· | ···················· | ···················· | ···················· | ···················· |
| E | 5568 | hCoV-19/Wales/PHWC-168445/2020\|EPI_ISL_494241\|2020 | ···················· | ···················· | ···················· | ···················· | ···················· | ···················· |
| E | 5569 | hCoV-19/Wales/PHWC-168454/2020\|EPI_ISL_494242\|2020 | ···················· | ···················· | ···················· | ···················· | ···················· | ···················· |
| E | 5570 | hCoV-19/Wales/PHWC-168463/2020\|EPI_ISL_494243\|2020 | ···················· | ···················· | ···················· | ···················· | ···················· | ···················· |
| E | 5571 | hCoV-19/Wales/PHWC-168472/2020\|EPI_ISL_494244\|2020 | ···················· | ···················· | ···················· | ···················· | ···················· | ···················· |
| E | 5572 | hCoV-19/Wales/PHWC-168481/2020\|EPI_ISL_494245\|2020 | ···················· | ···················· | ···················· | ···················· | ···················· | ···················· |
| E | 5573 | hCoV-19/Wales/PHWC-168506/2020\|EPI_ISL_494250\|2020 | ···················· | ···················· | ···················· | ···················· | ···················· | ···················· |
| E | 5574 | hCoV-19/Wales/PHWC-168515/2020\|EPI_ISL_494251\|2020 | ···················· | ···················· | ···················· | ···················· | ···················· | ···················· |
| E | 5575 | hCoV-19/Wales/PHWC-168542/2020\|EPI_ISL_494252\|2020 | ···················· | ···················· | ···················· | ···················· | ···················· | ···················· |
| E | 5576 | hCoV-19/Wales/PHWC-1685AC/2020\|EPI_ISL_494257\|2020 | ···················· | ···················· | ···················· | ···················· | ···················· | ···················· |
| E | 5577 | hCoV-19/Wales/PHWC-1685CA/2020\|EPI_ISL_494259\|2020 | ···················· | ···················· | ···················· | ···················· | ···················· | ···················· |
| E | 5578 | hCoV-19/Wales/PHWC-1685F7/2020\|EPI_ISL_494262\|2020 | ···················· | ···················· | ···················· | ···················· | ···················· | ···················· |
| E | 5579 | hCoV-19/Wales/PHWC-168603/2020\|EPI_ISL_494263\|2020 | ···················· | ···················· | ···················· | ···················· | ···················· | ···················· |
| E | 5580 | hCoV-19/Wales/PHWC-168621/2020\|EPI_ISL_494265\|2020 | ···················· | ···················· | ···················· | ···················· | ···················· | ···················· |
| E | 5581 | hCoV-19/Wales/PHWC-1686A9/2020\|EPI_ISL_494270\|2020 | ···················· | ···················· | ···················· | ···················· | ···················· | ···················· |
| E | 5582 | hCoV-19/Wales/PHWC-1686B8/2020\|EPI_ISL_494271\|2020 | ···················· | ···················· | ···················· | ···················· | ···················· | ···················· |
| E | 5583 | hCoV-19/Wales/PHWC-1686D6/2020\|EPI_ISL_494273\|2020 | ···················· | ···················· | ···················· | ···················· | ···················· | ···················· |
| E | 5584 | hCoV-19/Wales/PHWC-1686F4/2020\|EPI_ISL_494275\|2020 | ···················· | ···················· | ···················· | ···················· | ···················· | ···················· |
| E | 5585 | hCoV-19/Wales/PHWC-168788/2020\|EPI_ISL_494284\|2020 | ···················· | ···················· | ···················· | ···················· | ···················· | ···················· |
| E | 5586 | hCoV-19/Wales/PHWC-1687B5/2020\|EPI_ISL_494286\|2020 | ···················· | ···················· | ···················· | ···················· | ···················· | ···················· |
| E | 5587 | hCoV-19/Wales/PHWC-16880D/2020\|EPI_ISL_494291\|2020 | ···················· | ···················· | ···················· | ···················· | ···················· | ···················· |
| E | 5588 | hCoV-19/Wales/PHWC-168876/2020\|EPI_ISL_494296\|2020 | ···················· | ····A··············· | ···················· | ···················· | ···················· | ···················· |
| E | 5589 | hCoV-19/Wales/PHWC-1688A3/2020\|EPI_ISL_494299\|2020 | ···················· | ···················· | ···················· | ···················· | ···················· | ···················· |
| E | 5590 | hCoV-19/South Africa/KRISP-0417/2020\|EPI_ISL_49554 | ···················· | ···················· | ···················· | ···················· | ···················· | ···················· |
| E | 5591 | hCoV-19/South Africa/KRISP-0418/2020\|EPI_ISL_49554 | ···················· | ···················· | ···················· | ···················· | ···················· | ···················· |
| E | 5592 | hCoV-19/South Africa/KRISP-0420/2020\|EPI_ISL_49554 | ···················· | ···················· | ···················· | ···················· | ···················· | ···················· |
| E | 5593 | hCoV-19/South Africa/KRISP-0421/2020\|EPI_ISL_49554 | ···················· | ···················· | ···················· | ···················· | ···················· | ···················· |
| E | 5594 | hCoV-19/South Africa/KRISP-0426/2020\|EPI_ISL_49554 | ···················· | ···················· | ···················· | ···················· | ···················· | ···················· |
| E | 5595 | hCoV-19/South Africa/KRISP-0458/2020\|EPI_ISL_49554 | ···················· | ···················· | ···················· | ···················· | ···················· | ···················· |
| E | 5596 | hCoV-19/South Africa/KRISP-0460/2020\|EPI_ISL_49555 | ···················· | ···················· | ···················· | ···················· | ···················· | ···················· |
| E | 5597 | hCoV-19/South Africa/KRISP-0474/2020\|EPI_ISL_49555 | ···················· | ···················· | ···················· | ···················· | ···················· | ···················· |
| E | 5598 | hCoV-19/South Africa/KRISP-0475/2020\|EPI_ISL_49555 | ···················· | ···················· | ···················· | ···················· | ···················· | ···················· |
| E | 5599 | hCoV-19/South Africa/KRISP-0478/2020\|EPI_ISL_49555 | ···················· | ···················· | ···················· | ···················· | ···················· | ···················· |
| E | 5601 | hCoV-19/South Africa/KRISP-0487/2020\|EPI_ISL_49556 | ···················· | ···················· | ···················· | ···················· | ···················· | ···················· |
| E | 5602 | hCoV-19/USA/MI-UM-2006S000956/2020\|EPI_ISL_495563\| | ···················· | ···················· | ···················· | ···················· | ···················· | ···················· |
| E | 5603 | hCoV-19/USA/MI-UM-2006S000959/2020\|EPI_ISL_495564\| | ···················· | ···················· | ···················· | ···················· | ···················· | ···················· |
| E | 5604 | hCoV-19/USA/MI-UM-2006S001042/2020\|EPI_ISL_495566\| | ···················· | ···················· | ···················· | ···················· | ···················· | ···················· |
| E | 5605 | hCoV-19/USA/MI-UM-2006S001043/2020\|EPI_ISL_495567\| | ···················· | ···················· | ···················· | ···················· | ···················· | ···················· |
| E | 5607 | hCoV-19/USA/MI-UM-2006S001069/2020\|EPI_ISL_495572\| | ···················· | ···················· | ···················· | ···················· | ···················· | ···················· |
| E | 5608 | hCoV-19/USA/MI-UM-2006S001085/2020\|EPI_ISL_495574\| | ···················· | ···················· | ···················· | ···················· | ···················· | ···················· |
| E | 5609 | hCoV-19/USA/MI-UM-2006S001092/2020\|EPI_ISL_495576\| | ···················· | ···················· | ···················· | ···················· | ···················· | ···················· |
| E | 5610 | hCoV-19/USA/MI-UM-2006S001093/2020\|EPI_ISL_495577\| | ···················· | ···················· | ···················· | ···················· | ···················· | ···················· |
| E | 5612 | hCoV-19/USA/MI-UM-2006S001097/2020\|EPI_ISL_495579\| | ···················· | ···················· | ···················· | ···················· | ···················· | ···················· |
| E | 5613 | hCoV-19/USA/MI-UM-2006S001098/2020\|EPI_ISL_495580\| | ···················· | ···················· | ···················· | ···················· | ···················· | ···················· |
| E | 5614 | hCoV-19/USA/MI-UM-2006S001102/2020\|EPI_ISL_495581\| | ···················· | ···················· | ···················· | ···················· | ···················· | ···················· |
| E | 5616 | hCoV-19/USA/MI-UM-2006S001111/2020\|EPI_ISL_495583\| | ···················· | ···················· | ···················· | ···················· | ···················· | ···················· |
| E | 5618 | hCoV-19/USA/MI-UM-2006S001114/2020\|EPI_ISL_495585\| | ···················· | ···················· | ···················· | ···················· | ···················· | ···················· |
| E | 5619 | hCoV-19/USA/MI-UM-2006S001115/2020\|EPI_ISL_495586\| | ···················· | ···················· | ···················· | ···················· | ···················· | ···················· |
| E | 5621 | hCoV-19/USA/MI-UM-2006S001117/2020\|EPI_ISL_495588\| | ···················· | ···················· | ···················· | ···················· | ···················· | ···················· |
| E | 5624 | hCoV-19/USA/MI-UM-2006S001130/2020\|EPI_ISL_495591\| | ···················· | ···················· | ···················· | ···················· | ···················· | ···················· |
| E | 5625 | hCoV-19/USA/MI-UM-2006S001135/2020\|EPI_ISL_495592\| | ···················· | ···················· | ···················· | ···················· | ···················· | ···················· |
| E | 5626 | hCoV-19/USA/MI-UM-2006S001288/2020\|EPI_ISL_495593\| | ···················· | ···················· | ···················· | ···················· | ···················· | ···················· |
| E | 5627 | hCoV-19/USA/MI-UM-2006S005725/2020\|EPI_ISL_495594\| | ···················· | ···················· | ···················· | ···················· | ···················· | ···················· |
| E | 5628 | hCoV-19/USA/MI-UM-2006S005727/2020\|EPI_ISL_495595\| | ···················· | ···················· | ···················· | ···················· | ···················· | ···················· |
| E | 5630 | hCoV-19/USA/MN-MDH-1298/2020\|EPI_ISL_495597\|2020-0 | ···················· | ···················· | ···················· | ···················· | ···················· | ···················· |
| E | 5631 | hCoV-19/USA/MN-MDH-1299/2020\|EPI_ISL_495598\|2020-0 | ···················· | ···················· | ···················· | ···················· | ···················· | ···················· |
| E | 5632 | hCoV-19/USA/MN-MDH-1300/2020\|EPI_ISL_495599\|2020-0 | ···················· | ···················· | ···················· | ···················· | ···················· | ···················· |
| E | 5633 | hCoV-19/England/LOND-D681A/2020\|EPI_ISL_482200\|202 | ···················· | ···················· | ···················· | ···················· | ···················· | ···················· |
| E | 5634 | hCoV-19/England/LOND-D6188/2020\|EPI_ISL_482201\|202 | ···················· | ···················· | ···················· | ···················· | ···················· | ···················· |
| E | 5635 | hCoV-19/England/LOND-D615B/2020\|EPI_ISL_482202\|202 | ···················· | ···················· | ···················· | ···················· | ···················· | ···················· |
| E | 5636 | hCoV-19/England/LOND-D6285/2020\|EPI_ISL_482203\|202 | ···················· | ···················· | ···················· | ···················· | ···················· | ···················· |
| E | 5637 | hCoV-19/England/LOND-D62C1/2020\|EPI_ISL_482204\|202 | ···················· | ···················· | ···················· | ···················· | ···················· | ···················· |
| E | 5638 | hCoV-19/England/LOND-D62EF/2020\|EPI_ISL_482205\|202 | ···················· | ···················· | ···················· | ···················· | ···················· | ···················· |
| E | 5639 | hCoV-19/England/LOND-D621C/2020\|EPI_ISL_482206\|202 | ···················· | ···················· | ···················· | ···················· | ···················· | ···················· |
| E | 5640 | hCoV-19/England/LOND-D62D0/2020\|EPI_ISL_482208\|202 | ···················· | ···················· | ···················· | ···················· | ···················· | ···················· |
| E | 5641 | hCoV-19/England/LOND-D6777/2020\|EPI_ISL_482209\|202 | ···················· | ···················· | ···················· | ···················· | ···················· | ···················· |
| E | 5642 | hCoV-19/England/LOND-D67FF/2020\|EPI_ISL_482210\|202 | ···················· | ···················· | ···················· | ···················· | ···················· | ···················· |
| E | 5643 | hCoV-19/England/LOND-D6328/2020\|EPI_ISL_482211\|202 | ···················· | ···················· | ···················· | ···················· | ···················· | ···················· |
| E | 5644 | hCoV-19/England/LOND-D6795/2020\|EPI_ISL_482212\|202 | ···················· | ···················· | ···················· | ···················· | ···················· | ···················· |
| E | 5645 | hCoV-19/England/LOND-D6908/2020\|EPI_ISL_482213\|202 | ···················· | ···················· | ···················· | ···················· | ···················· | ···················· |
| E | 5646 | hCoV-19/England/LOND-D6258/2020\|EPI_ISL_482215\|202 | ···················· | ···················· | ···················· | ···················· | ···················· | ···················· |
| E | 5647 | hCoV-19/England/LOND-D614C/2020\|EPI_ISL_482217\|202 | ···················· | ···················· | ···················· | ···················· | ···················· | ···················· |
| E | 5648 | hCoV-19/USA/CA-ALSR-1072-SAN/2020\|EPI_ISL_483500\|2 | ···················· | ·A·················· | ···················· | ···················· | ···················· | ···················· |
| E | 5649 | hCoV-19/USA/CA-ALSR-1074-SAN/2020\|EPI_ISL_483501\|2 | ···················· | ·A·················· | ···················· | ···················· | ···················· | ···················· |
| E | 5650 | hCoV-19/USA/CA-ALSR-1075-SAN/2020\|EPI_ISL_483502\|2 | ···················· | ···················· | ···················· | ···················· | ···················· | ···················· |

| | | | | | | | | |
|---|---|---|---|---|---|---|---|---|
| E | 5651 | hCoV-19/USA/CA-ALSR-1076-SAN/2020\|EPI_ISL_483503\|2 | ···················· | ·A·················· | ···················· | ···················· | ···················· | ···················· |
| E | 5652 | hCoV-19/USA/CA-ALSR-1077-SAN/2020\|EPI_ISL_483504\|2 | ···················· | ···················· | ···················· | ···················· | ···················· | ···················· |
| E | 5653 | hCoV-19/USA/CA-ALSR-1078-SAN/2020\|EPI_ISL_483505\|2 | ···················· | ···················· | ···················· | ···················· | ···················· | ···················· |
| E | 5654 | hCoV-19/USA/CA-ALSR-1079-SAN/2020\|EPI_ISL_483506\|2 | ···················· | ···················· | ···················· | ···················· | ···················· | ···················· |
| E | 5655 | hCoV-19/USA/CA-ALSR-1080-SAN/2020\|EPI_ISL_483507\|2 | ···················· | ···················· | ···················· | ···················· | ···················· | ···················· |
| E | 5656 | hCoV-19/USA/CA-ALSR-1081-SAN/2020\|EPI_ISL_483508\|2 | ···················· | ·A·················· | ···················· | ···················· | ···················· | ···················· |
| E | 5657 | hCoV-19/USA/CA-ALSR-1082-SAN/2020\|EPI_ISL_483509\|2 | ···················· | ··C················· | ···················· | ···················· | ···················· | ···················· |
| E | 5658 | hCoV-19/USA/CA-ALSR-1083-SAN/2020\|EPI_ISL_483510\|2 | ···················· | ·A·················· | ···················· | ···················· | ···················· | ···················· |
| E | 5659 | hCoV-19/USA/CA-ALSR-1084-SAN/2020\|EPI_ISL_483511\|2 | ···················· | ·A·················· | ···················· | ···················· | ···················· | ···················· |
| E | 5660 | hCoV-19/USA/CA-ALSR-1085-SAN/2020\|EPI_ISL_483512\|2 | ···················· | ·A·················· | ···················· | ···················· | ···················· | ···················· |
| E | 5661 | hCoV-19/USA/CA-ALSR-1086-SAN/2020\|EPI_ISL_483513\|2 | ···················· | ···················· | ···················· | ···················· | ···················· | ···················· |
| E | 5662 | hCoV-19/USA/CA-ALSR-1087-SAN/2020\|EPI_ISL_483514\|2 | ·······T············ | ···················· | ···················· | ···················· | ···················· | ···················· |
| E | 5663 | hCoV-19/USA/CA-ALSR-1088-SAN/2020\|EPI_ISL_483515\|2 | ···················· | ·A·················· | ···················· | ···················· | ···················· | ···················· |
| E | 5664 | hCoV-19/USA/CA-ALSR-1090-SAN/2020\|EPI_ISL_483517\|2 | ···················· | ·A·················· | ···················· | ···················· | ···················· | ···················· |
| E | 5665 | hCoV-19/USA/CA-ALSR-1091-SAN/2020\|EPI_ISL_483518\|2 | ···················· | ·A·················· | ···················· | ···················· | ···················· | ···················· |
| E | 5666 | hCoV-19/USA/CA-ALSR-1095-SAN/2020\|EPI_ISL_483522\|2 | ···················· | ···················· | ···················· | ···················· | ···················· | ···················· |
| E | 5667 | hCoV-19/USA/CA-ALSR-1096-SAN/2020\|EPI_ISL_483523\|2 | ···················· | ···················· | ···················· | ···················· | ···················· | ···················· |
| E | 5668 | hCoV-19/USA/CA-ALSR-1097-SAN/2020\|EPI_ISL_483524\|2 | ···················· | ·A·················· | ···················· | ···················· | ···················· | ···················· |
| E | 5669 | hCoV-19/USA/CA-ALSR-1100-SAN/2020\|EPI_ISL_483526\|2 | ···················· | ···················· | ···················· | ···················· | ···················· | ···················· |
| E | 5670 | hCoV-19/USA/CA-ALSR-1101-SAN/2020\|EPI_ISL_483527\|2 | ···················· | ·A·················· | ···················· | ···················· | ···················· | ···················· |
| E | 5671 | hCoV-19/USA/CA-ALSR-1104-SAN/2020\|EPI_ISL_483529\|2 | ···················· | ·A·················· | ···················· | ···················· | ···················· | ···················· |
| E | 5672 | hCoV-19/USA/CA-ALSR-1106-SAN/2020\|EPI_ISL_483530\|2 | ···················· | ···················· | ···················· | ···················· | ···················· | ···················· |
| E | 5673 | hCoV-19/USA/CA-ALSR-1107-SAN/2020\|EPI_ISL_483531\|2 | ···················· | ·A·················· | ···················· | ···················· | ···················· | ···················· |
| E | 5674 | hCoV-19/USA/CA-ALSR-1110-SAN/2020\|EPI_ISL_483533\|2 | ···················· | ·A·················· | ···················· | ···················· | ···················· | ···················· |
| E | 5675 | hCoV-19/USA/CA-ALSR-1115-SAN/2020\|EPI_ISL_483538\|2 | ···················· | ·A·················· | ···················· | ···················· | ···················· | ···················· |
| E | 5676 | hCoV-19/USA/CA-ALSR-1117-SAN/2020\|EPI_ISL_483540\|2 | ···················· | ·A·················· | ···················· | ···················· | ···················· | ···················· |
| E | 5677 | hCoV-19/Bahrein/BAH-01/2020\|EPI_ISL_483542\|2020-03 | ···················· | ···················· | ···················· | ···················· | ···················· | ···················· |
| E | 5678 | hCoV-19/Bahrein/BAH-02/2020\|EPI_ISL_483543\|2020-03 | ···················· | ···················· | ···················· | ···················· | ···················· | ···················· |
| E | 5679 | hCoV-19/Bahrein/BAH-03/2020\|EPI_ISL_483544\|2020-03 | ···················· | ···················· | ···················· | ···················· | ···················· | ···················· |
| E | 5680 | hCoV-19/Bahrein/BAH-04/2020\|EPI_ISL_483545\|2020-03 | ···················· | ···················· | ···················· | ···················· | ···················· | ···················· |
| E | 5681 | hCoV-19/Bahrein/BAH-05/2020\|EPI_ISL_483546\|2020-03 | ···················· | ···················· | ···················· | ···················· | ···················· | ···················· |
| E | 5682 | hCoV-19/Bahrein/BAH-06/2020\|EPI_ISL_483547\|2020-03 | ···················· | ···················· | ···················· | ···················· | ···················· | ···················· |
| E | 5683 | hCoV-19/Bahrein/BAH-07/2020\|EPI_ISL_483548\|2020-03 | ···················· | ···················· | ···················· | ···················· | ···················· | ···················· |
| E | 5684 | hCoV-19/Bahrein/BAH-08/2020\|EPI_ISL_483549\|2020-03 | ···················· | ···················· | ···················· | ···················· | ···················· | ···················· |
| E | 5685 | hCoV-19/Bahrein/BAH-09/2020\|EPI_ISL_483550\|2020-03 | ···················· | ···················· | ···················· | ···················· | ···················· | ···················· |
| E | 5686 | hCoV-19/Bahrein/BAH-10/2020\|EPI_ISL_483551\|2020-03 | ···················· | ···················· | ···················· | ···················· | ···················· | ···················· |
| E | 5687 | hCoV-19/Bahrein/BAH-11/2020\|EPI_ISL_483552\|2020-03 | ···················· | ···················· | ···················· | ···················· | ···················· | ···················· |
| E | 5688 | hCoV-19/Bahrein/BAH-12/2020\|EPI_ISL_483553\|2020-03 | ···················· | ···················· | ···················· | ···················· | ···················· | ···················· |
| E | 5689 | hCoV-19/Bahrein/BAH-13/2020\|EPI_ISL_483554\|2020-03 | ···················· | ···················· | ···················· | ···················· | ···················· | ···················· |
| E | 5690 | hCoV-19/Bahrein/BAH-14/2020\|EPI_ISL_483555\|2020-04 | ···················· | ···················· | ···················· | ···················· | ···················· | ···················· |
| E | 5691 | hCoV-19/Bahrein/BAH-15/2020\|EPI_ISL_483556\|2020-04 | ···················· | ···················· | ···················· | ···················· | ···················· | ···················· |
| E | 5692 | hCoV-19/Bahrein/BAH-16/2020\|EPI_ISL_483557\|2020-04 | ···················· | ···················· | ···················· | ···················· | ···················· | ···················· |
| E | 5693 | hCoV-19/Bahrein/BAH-17/2020\|EPI_ISL_483558\|2020-04 | ···················· | ···················· | ···················· | ···················· | ···················· | ···················· |
| E | 5694 | hCoV-19/Bahrein/BAH-18/2020\|EPI_ISL_483559\|2020-04 | ···················· | ···················· | ···················· | ···················· | ···················· | ···················· |
| E | 5695 | hCoV-19/USA/MI-UM-2006S001122/2020\|EPI_ISL_484806\| | ···················· | ···················· | ···················· | ···················· | ···················· | ···················· |
| E | 5696 | hCoV-19/USA/WI-UW-432/2020\|EPI_ISL_484807\|2020-06- | ···················· | ···················· | ···················· | ···················· | ···················· | ···················· |
| E | 5697 | hCoV-19/USA/WI-UW-433/2020\|EPI_ISL_484808\|2020-06- | ···················· | ···················· | ···················· | ···················· | ···················· | ···················· |
| E | 5698 | hCoV-19/USA/WI-UW-434/2020\|EPI_ISL_484809\|2020-06- | ···················· | ···················· | ···················· | ···················· | ···················· | ···················· |
| E | 5699 | hCoV-19/USA/WI-UW-435/2020\|EPI_ISL_484810\|2020-06- | ···················· | ···················· | ···················· | ···················· | ···················· | ···················· |
| E | 5700 | hCoV-19/USA/WI-UW-436/2020\|EPI_ISL_484811\|2020-06- | ···················· | ···················· | ···················· | ···················· | ···················· | ···················· |
| E | 5701 | hCoV-19/USA/WI-UW-437/2020\|EPI_ISL_484812\|2020-06- | ·T·················· | ···················· | ···················· | ···················· | ···················· | ···················· |
| E | 5702 | hCoV-19/USA/WI-UW-438/2020\|EPI_ISL_484813\|2020-06- | ···················· | ···················· | ···················· | ···················· | ···················· | ···················· |
| E | 5703 | hCoV-19/USA/WI-UW-439/2020\|EPI_ISL_484814\|2020-06- | ···················· | ···················· | ···················· | ···················· | ···················· | ···················· |
| E | 5704 | hCoV-19/USA/WI-UW-440/2020\|EPI_ISL_484815\|2020-06- | ···················· | ···················· | ···················· | ···················· | ···················· | ···················· |
| E | 5705 | hCoV-19/USA/WI-UW-442/2020\|EPI_ISL_484817\|2020-06- | ···················· | ···················· | ···················· | ···················· | ···················· | ···················· |
| E | 5706 | hCoV-19/USA/WI-UW-443/2020\|EPI_ISL_484818\|2020-06- | ···················· | ···················· | ···················· | ···················· | ···················· | ···················· |
| E | 5707 | hCoV-19/USA/WI-UW-444/2020\|EPI_ISL_484819\|2020-06- | ···················· | ···················· | ···················· | ···················· | ···················· | ···················· |
| E | 5708 | hCoV-19/USA/WI-UW-445/2020\|EPI_ISL_484820\|2020-06- | ···················· | ···················· | ···················· | ···················· | ···················· | ···················· |
| E | 5709 | hCoV-19/USA/WI-UW-446/2020\|EPI_ISL_484821\|2020-06- | ···················· | ···················· | ···················· | ···················· | ···················· | ···················· |
| E | 5710 | hCoV-19/USA/WI-UW-447/2020\|EPI_ISL_484822\|2020-06- | ···················· | ···················· | ···················· | ···················· | ···················· | ···················· |
| E | 5711 | hCoV-19/USA/WI-UW-448/2020\|EPI_ISL_484823\|2020-06- | ···················· | ···················· | ···················· | ···················· | ···················· | ···················· |
| E | 5712 | hCoV-19/USA/WI-UW-449/2020\|EPI_ISL_484824\|2020-06- | ···················· | ···················· | ···················· | ···················· | ···················· | ···················· |
| E | 5713 | hCoV-19/USA/WI-UW-450/2020\|EPI_ISL_484825\|2020-06- | ···················· | ···················· | ···················· | ···················· | ···················· | ···················· |
| E | 5714 | hCoV-19/USA/WI-UW-451/2020\|EPI_ISL_484826\|2020-06- | ···················· | ···················· | ···················· | ···················· | ···················· | ···················· |
| E | 5715 | hCoV-19/USA/WI-UW-452/2020\|EPI_ISL_484827\|2020-06- | ···················· | ···················· | ···················· | ···················· | ···················· | ···················· |
| E | 5716 | hCoV-19/USA/WI-UW-453/2020\|EPI_ISL_484828\|2020-06- | ···················· | ···················· | ···················· | ···················· | ···················· | ···················· |
| E | 5717 | hCoV-19/USA/WI-UW-455/2020\|EPI_ISL_484830\|2020-06- | ·T·················· | ···················· | ···················· | ···················· | ···················· | ···················· |
| E | 5718 | hCoV-19/USA/WI-UW-457/2020\|EPI_ISL_484832\|2020-06- | ·T·················· | ···················· | ···················· | ···················· | ···················· | ···················· |
| E | 5719 | hCoV-19/USA/WI-UW-458/2020\|EPI_ISL_484833\|2020-06- | ···················· | ···················· | ···················· | ···················· | ···················· | ···················· |
| E | 5720 | hCoV-19/USA/WI-UW-459/2020\|EPI_ISL_484834\|2020-06- | ···················· | ···················· | ···················· | ···················· | ···················· | ···················· |
| E | 5721 | hCoV-19/USA/WI-UW-460/2020\|EPI_ISL_484835\|2020-06- | ···················· | ···················· | ···················· | ···················· | ···················· | ···················· |
| E | 5722 | hCoV-19/USA/WI-UW-461/2020\|EPI_ISL_484836\|2020-06- | ···················· | ···················· | ···················· | ···················· | ···················· | ···················· |
| E | 5723 | hCoV-19/USA/WI-UW-462/2020\|EPI_ISL_484837\|2020-06- | ···················· | ···················· | ···················· | ···················· | ···················· | ···················· |
| E | 5724 | hCoV-19/USA/WI-UW-463/2020\|EPI_ISL_484838\|2020-06- | ···················· | ···················· | ···················· | ···················· | ···················· | ···················· |
| E | 5725 | hCoV-19/USA/WI-UW-464/2020\|EPI_ISL_484839\|2020-06- | ···················· | ···················· | ···················· | ···················· | ···················· | ···················· |
| E | 5726 | hCoV-19/USA/WI-UW-465/2020\|EPI_ISL_484840\|2020-06- | ···················· | ···················· | ···················· | ···················· | ···················· | ···················· |
| E | 5727 | hCoV-19/USA/WI-UW-466/2020\|EPI_ISL_484841\|2020-06- | ···················· | ···················· | ···················· | ···················· | ···················· | ···················· |
| E | 5728 | hCoV-19/USA/WI-UW-467/2020\|EPI_ISL_484842\|2020-06- | ···················· | ···················· | ···················· | ···················· | ···················· | ···················· |
| E | 5729 | hCoV-19/USA/WI-UW-468/2020\|EPI_ISL_484843\|2020-06- | ···················· | ···················· | ···················· | ···················· | ···················· | ···················· |
| E | 5730 | hCoV-19/USA/WI-UW-469/2020\|EPI_ISL_484844\|2020-06- | ···················· | ···················· | ···················· | ···················· | ···················· | ···················· |

| | | | | | | | |
|---|---|---|---|---|---|---|---|
| E | 5731 hCoV-19/USA/WI-UW-470/2020\|EPI_ISL_484845\|2020-06- | ···················· | ···················· | ····················· | ···················· | ···················· | ···················· |
| E | 5732 hCoV-19/USA/WI-UW-471/2020\|EPI_ISL_484846\|2020-06- | ···················· | ···················· | ····················· | ···················· | ···················· | ···················· |
| E | 5733 hCoV-19/USA/IL-UW-472/2020\|EPI_ISL_484847\|2020-06- | ···················· | ···················· | ····················· | ···················· | ···················· | ···················· |
| E | 5734 hCoV-19/USA/WI-UW-474/2020\|EPI_ISL_484849\|2020-06- | ···················· | ···················· | ····················· | ···················· | ···················· | ···················· |
| E | 5735 hCoV-19/USA/WI-UW-476/2020\|EPI_ISL_484851\|2020-06- | ···················· | ···················· | ····················· | ···················· | ···················· | ···················· |
| E | 5736 hCoV-19/USA/WI-UW-477/2020\|EPI_ISL_484852\|2020-06- | ···················· | ···················· | ····················· | ···················· | ···················· | ···················· |
| E | 5737 hCoV-19/USA/WI-UW-482/2020\|EPI_ISL_484857\|2020-06- | ···················· | ···················· | ····················· | ···················· | ···················· | ···················· |
| E | 5738 hCoV-19/USA/WI-UW-483/2020\|EPI_ISL_484858\|2020-06- | ···················· | ···················· | ····················· | ···················· | ···················· | ···················· |
| E | 5739 hCoV-19/USA/WI-UW-485/2020\|EPI_ISL_484860\|2020-06- | ···················· | ···················· | ····················· | ···················· | ···················· | ···················· |
| E | 5740 hCoV-19/USA/WI-UW-486/2020\|EPI_ISL_484861\|2020-06- | ···················· | ···················· | ····················· | ···················· | ···················· | ···················· |
| E | 5741 hCoV-19/USA/IL-UW-487/2020\|EPI_ISL_484862\|2020-06- | ···················· | ···················· | ····················· | ···················· | ···················· | ···················· |
| E | 5742 hCoV-19/USA/WI-UW-488/2020\|EPI_ISL_484863\|2020-06- | ···················· | ···················· | ····················· | ···················· | ···················· | ···················· |
| E | 5743 hCoV-19/USA/WI-UW-489/2020\|EPI_ISL_484864\|2020-06- | ···················· | ···················· | ····················· | ···················· | ···················· | ···················· |
| E | 5744 hCoV-19/USA/WI-UW-490/2020\|EPI_ISL_484865\|2020-06- | ···················· | ···················· | ····················· | ···················· | ···················· | ···················· |
| E | 5745 hCoV-19/USA/WI-UW-491/2020\|EPI_ISL_484866\|2020-06- | ···················· | ···················· | ····················· | ···················· | ···················· | ···················· |
| E | 5746 hCoV-19/USA/WI-UW-492/2020\|EPI_ISL_484867\|2020-06- | ···················· | ···················· | ····················· | ···················· | ···················· | ···················· |
| E | 5747 hCoV-19/USA/WI-UW-493/2020\|EPI_ISL_484868\|2020-06- | ···················· | ···················· | ····················· | ···················· | ···················· | ···················· |
| E | 5748 hCoV-19/USA/WI-UW-494/2020\|EPI_ISL_484869\|2020-06- | ···················· | ···················· | ····················· | ···················· | ···················· | ···················· |
| E | 5749 hCoV-19/USA/WI-UW-495/2020\|EPI_ISL_484870\|2020-06- | ···················· | ···················· | ····················· | ···················· | ···················· | ···················· |
| E | 5750 hCoV-19/USA/WI-UW-496/2020\|EPI_ISL_484871\|2020-06- | ···················· | ···················· | ····················· | ···················· | ···················· | ···················· |
| E | 5751 hCoV-19/USA/WI-UW-497/2020\|EPI_ISL_484872\|2020-06- | ···················· | ···················· | ····················· | ···················· | ···················· | ···················· |
| E | 5752 hCoV-19/USA/WI-UW-498/2020\|EPI_ISL_484873\|2020-06- | ···················· | ···················· | ····················· | ···················· | ···················· | ···················· |
| E | 5753 hCoV-19/USA/WI-UW-499/2020\|EPI_ISL_484874\|2020-06- | ···················· | ···················· | ····················· | ···················· | ···················· | ···················· |
| E | 5754 hCoV-19/USA/WI-UW-500/2020\|EPI_ISL_484875\|2020-06- | ···················· | ···················· | ····················· | ···················· | ···················· | ···················· |
| E | 5755 hCoV-19/USA/WI-UW-501/2020\|EPI_ISL_484876\|2020-06- | ···················· | ···················· | ····················· | ···················· | ···················· | ···················· |
| E | 5756 hCoV-19/USA/WI-UW-502/2020\|EPI_ISL_484877\|2020-06- | ···················· | ···················· | ····················· | ···················· | ···················· | ···················· |
| E | 5757 hCoV-19/USA/IL-UW-503/2020\|EPI_ISL_484878\|2020-05- | ···················· | ···················· | ····················· | ···················· | ···················· | ···················· |
| E | 5758 hCoV-19/USA/WI-UW-504/2020\|EPI_ISL_484879\|2020-06- | ···················· | ···················· | ····················· | ···················· | ···················· | ···················· |
| E | 5759 hCoV-19/USA/WI-UW-505/2020\|EPI_ISL_484880\|2020-06- | ···················· | ···················· | ····················· | ···················· | ···················· | ···················· |
| E | 5760 hCoV-19/USA/IL-UW-506/2020\|EPI_ISL_484881\|2020-06- | ···················· | ···················· | ····················· | ···················· | ···················· | ···················· |
| E | 5761 hCoV-19/USA/IL-UW-507/2020\|EPI_ISL_484882\|2020-05- | ···················· | ···················· | ····················· | ···················· | ···················· | ···················· |
| E | 5762 hCoV-19/USA/WI-UW-508/2020\|EPI_ISL_484883\|2020-06- | ···················· | ···················· | ····················· | ···················· | ···················· | ···················· |
| E | 5763 hCoV-19/USA/WI-UW-509/2020\|EPI_ISL_484884\|2020-06- | ···················· | ···················· | ····················· | ···················· | ···················· | ···················· |
| E | 5764 hCoV-19/USA/CT-UW-510/2020\|EPI_ISL_484885\|2020-06- | ···················· | ···················· | ····················· | ···················· | ···················· | ···················· |
| E | 5765 hCoV-19/USA/WI-UW-511/2020\|EPI_ISL_484886\|2020-06- | ···················· | ···················· | ····················· | ···················· | ···················· | ···················· |
| E | 5766 hCoV-19/USA/WI-UW-513/2020\|EPI_ISL_484888\|2020-06- | ···················· | ···················· | ····················· | ···················· | ···················· | ···················· |
| E | 5767 hCoV-19/USA/WI-UW-514/2020\|EPI_ISL_484889\|2020-06- | ···················· | ···················· | ····················· | ···················· | ···················· | ···················· |
| E | 5768 hCoV-19/USA/IL-UW-515/2020\|EPI_ISL_484890\|2020-06- | ···················· | ···················· | ····················· | ···················· | ···················· | ···················· |
| E | 5769 hCoV-19/USA/WI-UW-517/2020\|EPI_ISL_484892\|2020-06- | ···················· | ···················· | ····················· | ···················· | ···················· | ···················· |
| E | 5770 hCoV-19/Nigeria/OY233-CV66/2020\|EPI_ISL_487091\|202 | ···················· | ···················· | ····················· | ···················· | ···················· | ···················· |
| E | 5771 hCoV-19/Nigeria/OY094-CV57/2020\|EPI_ISL_487099\|202 | ···················· | ···················· | ····················· | ···················· | ···················· | ···················· |
| E | 5772 hCoV-19/England/NORT-297B1A/2020\|EPI_ISL_488101\|20 | ···················· | ···················· | ····················· | ···················· | ···················· | ···················· |
| E | 5773 hCoV-19/England/NORT-29702A/2020\|EPI_ISL_488102\|20 | ···················· | ···················· | ····················· | ···················· | ···················· | ···················· |
| E | 5774 hCoV-19/England/NORT-29EABC/2020\|EPI_ISL_488103\|20 | ···················· | ···················· | ····················· | ···················· | ···················· | ···················· |
| E | 5775 hCoV-19/England/NORT-29D507/2020\|EPI_ISL_488104\|20 | ···················· | ···················· | ····················· | ···················· | ···················· | ···················· |
| E | 5776 hCoV-19/England/NORT-29DA44/2020\|EPI_ISL_488105\|20 | ···················· | ···················· | ····················· | ···················· | ···················· | ···················· |
| E | 5777 hCoV-19/England/NORT-297D32/2020\|EPI_ISL_488106\|20 | ···················· | ···················· | ····················· | ···················· | ···················· | ···················· |
| E | 5778 hCoV-19/England/NORT-297DBA/2020\|EPI_ISL_488107\|20 | ···················· | ···················· | ····················· | ···················· | ······K············· | ···················· |
| E | 5779 hCoV-19/England/NORT-296D42/2020\|EPI_ISL_488108\|20 | ···················· | ···················· | ····················· | ···················· | ···················· | ···················· |
| E | 5780 hCoV-19/England/NORT-297ED5/2020\|EPI_ISL_488110\|20 | ···················· | ···················· | ····················· | ···················· | ···················· | ···················· |
| E | 5781 hCoV-19/England/NORT-29F189/2020\|EPI_ISL_488111\|20 | ···················· | ···················· | ····················· | ···················· | ···················· | ···················· |
| E | 5782 hCoV-19/England/NORT-29EE38/2020\|EPI_ISL_488113\|20 | ···················· | ···················· | ····················· | ···················· | ···················· | ···················· |
| E | 5783 hCoV-19/England/NORT-297A59/2020\|EPI_ISL_488114\|20 | ···················· | ···················· | ····················· | ···················· | ···················· | ···················· |
| E | 5784 hCoV-19/England/NORT-29EADA/2020\|EPI_ISL_488115\|20 | ···················· | ···················· | ····················· | ···················· | ···················· | ···················· |
| E | 5785 hCoV-19/England/NORT-29D3EF/2020\|EPI_ISL_488116\|20 | ···················· | ···················· | ····················· | ···················· | ···················· | ···················· |
| E | 5786 hCoV-19/England/NORT-297163/2020\|EPI_ISL_488117\|20 | ···················· | ···················· | ····················· | ···················· | ···················· | ···················· |
| E | 5787 hCoV-19/England/NORT-29E005/2020\|EPI_ISL_488118\|20 | ···················· | ···················· | ····················· | ···················· | ···················· | ···················· |
| E | 5788 hCoV-19/England/NORT-296BDF/2020\|EPI_ISL_488119\|20 | ···················· | ···················· | ····················· | ···················· | ···················· | ···················· |
| E | 5789 hCoV-19/England/NORT-296B84/2020\|EPI_ISL_488120\|20 | ···················· | ···················· | ····················· | ···················· | ···················· | ···················· |
| E | 5790 hCoV-19/England/NORT-297075/2020\|EPI_ISL_488123\|20 | ···················· | ···················· | ····················· | ···················· | ···················· | ···················· |
| E | 5791 hCoV-19/England/NORT-29D3C1/2020\|EPI_ISL_488124\|20 | ···················· | ···················· | ····················· | ···················· | ···················· | ···················· |
| E | 5792 hCoV-19/England/NORT-29D947/2020\|EPI_ISL_488126\|20 | ···················· | ···················· | ····················· | ···················· | ···················· | ···················· |
| E | 5793 hCoV-19/England/NORT-29EC5C/2020\|EPI_ISL_488134\|20 | ···················· | ···················· | ····················· | ···················· | ···················· | ···················· |
| E | 5794 hCoV-19/England/BRIS-12E627/2020\|EPI_ISL_488380\|20 | ···················· | ···················· | ····················· | ···················· | ···················· | ···················· |
| E | 5795 hCoV-19/England/BRIS-12E584/2020\|EPI_ISL_488381\|20 | ···················· | ···················· | ····················· | ···················· | ···················· | ···················· |
| E | 5796 hCoV-19/England/BRIS-12FB54/2020\|EPI_ISL_488382\|20 | ···················· | ···················· | ····················· | ···················· | ···················· | ···················· |
| E | 5797 hCoV-19/England/BRIS-12EA58/2020\|EPI_ISL_488383\|20 | ···················· | ···················· | ····················· | ···················· | ···················· | ···················· |
| E | 5798 hCoV-19/England/BRIS-12E618/2020\|EPI_ISL_488384\|20 | ···················· | ···················· | ····················· | ···················· | ···················· | ···················· |
| E | 5799 hCoV-19/England/BRIS-12F134/2020\|EPI_ISL_488385\|20 | ···················· | ···················· | ····················· | ···················· | ···················· | ···················· |
| E | 5800 hCoV-19/England/BRIS-12F301/2020\|EPI_ISL_488386\|20 | ···················· | ···················· | ····················· | ···················· | ···················· | ···················· |
| E | 5801 hCoV-19/England/BRIS-12F565/2020\|EPI_ISL_488388\|20 | ···················· | ···················· | ····················· | ···················· | ···················· | ···················· |
| E | 5802 hCoV-19/England/BRIS-12FAC0/2020\|EPI_ISL_488389\|20 | ···················· | ···················· | ····················· | ···················· | ···················· | ···················· |
| E | 5803 hCoV-19/England/BRIS-12FAFD/2020\|EPI_ISL_488390\|20 | ···················· | ···················· | ····················· | ···················· | ···················· | ···················· |
| E | 5804 hCoV-19/England/BRIS-12FB09/2020\|EPI_ISL_488392\|20 | ···················· | ···················· | ····················· | ···················· | ···················· | ···················· |
| E | 5805 hCoV-19/England/BRIS-12E79D/2020\|EPI_ISL_488395\|20 | ···················· | ···················· | ····················· | ···················· | ···················· | ···················· |
| E | 5806 hCoV-19/England/BRIS-12F019/2020\|EPI_ISL_488398\|20 | ···················· | ···················· | ····················· | ···················· | ···················· | ···················· |
| E | 5807 hCoV-19/England/CAMB-1ACA16/2020\|EPI_ISL_489427\|20 | ···················· | ···················· | ····················· | ···················· | ···················· | ···················· |
| E | 5808 hCoV-19/England/CAMB-1AC4AF/2020\|EPI_ISL_489428\|20 | ···················· | ···················· | ····················· | ···················· | ···················· | ···················· |
| E | 5809 hCoV-19/Scotland/GCVR-171A65/2020\|EPI_ISL_489429\|2 | ···················· | ···················· | ····················· | ···················· | ···················· | ···················· |
| E | 5810 hCoV-19/England/CAMB-1AC68B/2020\|EPI_ISL_489430\|20 | ···················· | ···················· | ····················· | ···················· | ···················· | ···················· |

| | | | | | | | | |
|---|---|---|---|---|---|---|---|---|
| E | 5811 | hCoV-19/England/CAMB-1AC5AC/2020\|EPI_ISL_489431\|20 | ···················· | ···················· | ···················· | ···················· | ···················· | ···················· |
| E | 5812 | hCoV-19/England/CAMB-1AC5E8/2020\|EPI_ISL_489432\|20 | ···················· | ···················· | ···················· | ···················· | ···················· | ···················· |
| E | 5813 | hCoV-19/England/CAMB-1AD699/2020\|EPI_ISL_489435\|20 | ···················· | ···················· | ···················· | ···················· | ···················· | ···················· |
| E | 5814 | hCoV-19/England/CAMB-1ACFAE/2020\|EPI_ISL_489436\|20 | ···················· | ···················· | ···················· | ···················· | ···················· | ···················· |
| E | 5815 | hCoV-19/England/CAMB-1ACE38/2020\|EPI_ISL_489437\|20 | ···················· | ···················· | ···················· | ···················· | ···················· | ···················· |
| E | 5816 | hCoV-19/England/CAMB-1AC463/2020\|EPI_ISL_489438\|20 | ···················· | ···················· | ···················· | ···················· | ···················· | ···················· |
| E | 5817 | hCoV-19/Scotland/GCVR-17192C/2020\|EPI_ISL_489442\|2 | ···················· | ···················· | ···················· | ···················· | ···················· | ···················· |
| E | 5818 | hCoV-19/England/CAMB-1AC58E/2020\|EPI_ISL_489444\|20 | ···················· | ···················· | ···················· | ···················· | ···················· | ···················· |
| E | 5819 | hCoV-19/England/CAMB-1AD56F/2020\|EPI_ISL_489447\|20 | ···················· | ···················· | ···················· | ···················· | ···················· | ···················· |
| E | 5820 | hCoV-19/England/CAMB-1AC4BE/2020\|EPI_ISL_489448\|20 | ···················· | ···················· | ···················· | ···················· | ···················· | ···················· |
| E | 5821 | hCoV-19/Scotland/GCVR-1719E0/2020\|EPI_ISL_489449\|2 | ···················· | ···················· | ···················· | ···················· | ···················· | ···················· |
| E | 5822 | hCoV-19/England/CAMB-1ACC10/2020\|EPI_ISL_489451\|20 | ···················· | ···················· | ···················· | ···················· | ···················· | ···················· |
| E | 5823 | hCoV-19/England/CAMB-1AC80D/2020\|EPI_ISL_489452\|20 | ···················· | ···················· | ···················· | ···················· | ···················· | ···················· |
| E | 5824 | hCoV-19/England/CAMB-1AD3FC/2020\|EPI_ISL_489453\|20 | ···················· | ···················· | ···················· | ···················· | ···················· | ···················· |
| E | 5825 | hCoV-19/England/CAMB-1AC3DF/2020\|EPI_ISL_489454\|20 | ···················· | ···················· | ···················· | ···················· | ···················· | ···················· |
| E | 5826 | hCoV-19/Scotland/GCVR-171A29/2020\|EPI_ISL_489455\|2 | ···················· | ·A·················· | ···················· | ···················· | ···················· | ···················· |
| E | 5827 | hCoV-19/England/CAMB-1AC69A/2020\|EPI_ISL_489456\|20 | ···················· | ···················· | ···················· | ···················· | ···················· | ···················· |
| E | 5828 | hCoV-19/England/CAMB-1ACE29/2020\|EPI_ISL_489457\|20 | ···················· | ···················· | ···················· | ···················· | ···················· | ···················· |
| E | 5829 | hCoV-19/England/CAMB-1ACD4A/2020\|EPI_ISL_489467\|20 | ···················· | ···················· | ···················· | ···················· | ···················· | ···················· |
| E | 5830 | hCoV-19/England/CAMB-1AD4F9/2020\|EPI_ISL_489468\|20 | ···················· | ···················· | ···················· | ···················· | ···················· | ···················· |
| E | 5831 | hCoV-19/England/LIVE-A4235/2020\|EPI_ISL_490410\|202 | ···················· | ···················· | ···················· | ···················· | ···················· | ···················· |
| E | 5832 | hCoV-19/England/LIVE-A42F9/2020\|EPI_ISL_490412\|202 | ···················· | ···················· | ···················· | ···················· | ···················· | ···················· |
| E | 5833 | hCoV-19/England/LIVE-A4314/2020\|EPI_ISL_490413\|202 | ···················· | ···················· | ···················· | ···················· | ···················· | ···················· |
| E | 5834 | hCoV-19/England/LIVE-A4323/2020\|EPI_ISL_490414\|202 | ···················· | ···················· | ···················· | ···················· | ···················· | ···················· |
| E | 5835 | hCoV-19/England/LIVE-A4332/2020\|EPI_ISL_490415\|202 | ···················· | ···················· | ···················· | ···················· | ···················· | ···················· |
| E | 5836 | hCoV-19/England/LIVE-A436F/2020\|EPI_ISL_490417\|202 | ···················· | ···················· | ···················· | ···················· | ···················· | ···················· |
| E | 5837 | hCoV-19/England/LIVE-A437E/2020\|EPI_ISL_490418\|202 | ···················· | ···················· | ···················· | ···················· | ···················· | ···················· |
| E | 5838 | hCoV-19/England/LIVE-A438D/2020\|EPI_ISL_490419\|202 | ···················· | ···················· | ···················· | ···················· | ···················· | ···················· |
| E | 5839 | hCoV-19/England/LIVE-A43AB/2020\|EPI_ISL_490420\|202 | ···················· | ···················· | ···················· | ···················· | ···················· | ···················· |
| E | 5840 | hCoV-19/England/LOND-124C67A/2020\|EPI_ISL_490421\|2 | ···················· | ···················· | ···················· | ···················· | ···················· | ···················· |
| E | 5841 | hCoV-19/England/LOND-124C698/2020\|EPI_ISL_490422\|2 | ···················· | ···················· | ···················· | ···················· | ···················· | ···················· |
| E | 5842 | hCoV-19/England/LOND-124C6B6/2020\|EPI_ISL_490423\|2 | ···················· | ···················· | ···················· | ···················· | ···················· | ···················· |
| E | 5843 | hCoV-19/England/LOND-124C6E3/2020\|EPI_ISL_490424\|2 | ···················· | ···················· | ···················· | ···················· | ···················· | ···················· |
| E | 5844 | hCoV-19/England/LOND-124C6F2/2020\|EPI_ISL_490425\|2 | ···················· | ···················· | ···················· | ···················· | ···················· | ···················· |
| E | 5845 | hCoV-19/England/LOND-124C70E/2020\|EPI_ISL_490426\|2 | ···················· | ···················· | ···················· | ···················· | ···················· | ···················· |
| E | 5846 | hCoV-19/England/LOND-124C73B/2020\|EPI_ISL_490427\|2 | ···················· | ···················· | ···················· | ···················· | ···················· | ···················· |
| E | 5847 | hCoV-19/England/LOND-124C768/2020\|EPI_ISL_490428\|2 | ···················· | ·A·················· | ···················· | ···················· | ···················· | ···················· |
| E | 5848 | hCoV-19/England/LOND-124C777/2020\|EPI_ISL_490429\|2 | ···················· | ···················· | ···················· | ···················· | ···················· | ···················· |
| E | 5849 | hCoV-19/England/LOND-124C7A4/2020\|EPI_ISL_490430\|2 | ···················· | ···················· | ···················· | ···················· | ···················· | ···················· |
| E | 5850 | hCoV-19/England/LOND-124C7B3/2020\|EPI_ISL_490431\|2 | ···················· | ···················· | ···················· | ···················· | ···················· | ···················· |
| E | 5851 | hCoV-19/England/LOND-124C7D1/2020\|EPI_ISL_490432\|2 | ···················· | ···················· | ···················· | ···················· | ···················· | ···················· |
| E | 5852 | hCoV-19/England/LOND-124C7FF/2020\|EPI_ISL_490433\|2 | ···················· | ···················· | ···················· | ···················· | ···················· | ···················· |
| E | 5853 | hCoV-19/England/LOND-124C80B/2020\|EPI_ISL_490434\|2 | ···················· | ···················· | ···················· | ···················· | ···················· | ···················· |
| E | 5854 | hCoV-19/England/LOND-124C81A/2020\|EPI_ISL_490435\|2 | ···················· | ···················· | ···················· | ···················· | ···················· | ···················· |
| E | 5855 | hCoV-19/England/NORT-29310D/2020\|EPI_ISL_490444\|20 | ···················· | ···················· | ···················· | ···················· | ···················· | ···················· |
| E | 5856 | hCoV-19/England/NORT-2948DA/2020\|EPI_ISL_490449\|20 | ···················· | ···················· | ···················· | ···················· | ···················· | ···················· |
| E | 5857 | hCoV-19/Scotland/CVR2522/2020\|EPI_ISL_490690\|2020- | ···················· | ···················· | ···················· | ···················· | ···················· | ···················· |
| E | 5859 | hCoV-19/Scotland/CVR3506/2020\|EPI_ISL_490694\|2020- | ···················· | ···················· | ···················· | ···················· | ···················· | ···················· |
| E | 5860 | hCoV-19/Scotland/CVR3899/2020\|EPI_ISL_490695\|2020- | ···················· | ···················· | ···················· | ···················· | ···················· | ···················· |
| E | 5862 | hCoV-19/Scotland/CVR3901/2020\|EPI_ISL_490697\|2020- | ···················· | ···················· | ···················· | ···················· | ···················· | ···················· |
| E | 5863 | hCoV-19/Scotland/CVR3902/2020\|EPI_ISL_490698\|2020- | ···················· | ·A·················· | ···················· | ···················· | ···················· | ···················· |
| E | 5864 | hCoV-19/Wales/PHWC-1680D8/2020\|EPI_ISL_494200\|2020 | ···················· | ···················· | ···················· | ···················· | ···················· | ···················· |
| E | 5865 | hCoV-19/Wales/PHWC-1680E7/2020\|EPI_ISL_494201\|2020 | ···················· | ···················· | ···················· | ···················· | ···················· | ···················· |
| E | 5866 | hCoV-19/Wales/PHWC-1680F6/2020\|EPI_ISL_494202\|2020 | ···················· | ···················· | ···················· | ···················· | ···················· | ···················· |
| E | 5867 | hCoV-19/Wales/PHWC-1681A8/2020\|EPI_ISL_494208\|2020 | ···················· | ···················· | ···················· | ···················· | ···················· | ···················· |
| E | 5868 | hCoV-19/Wales/PHWC-1681B7/2020\|EPI_ISL_494209\|2020 | ···················· | ···················· | ···················· | ···················· | ···················· | ···················· |
| E | 5869 | hCoV-19/USA/WI-UW-796/2020\|EPI_ISL_495500\|2020-07- | ···················· | ···················· | ···················· | ···················· | ···················· | ···················· |
| E | 5870 | hCoV-19/USA/WI-UW-797/2020\|EPI_ISL_495501\|2020-07- | ···················· | ···················· | ···················· | ···················· | ···················· | ···················· |
| E | 5871 | hCoV-19/USA/WI-UW-800/2020\|EPI_ISL_495504\|2020-07- | ···················· | ···················· | ···················· | ···················· | ···················· | ···················· |
| E | 5872 | hCoV-19/USA/WI-UW-801/2020\|EPI_ISL_495505\|2020-07- | ···················· | ···················· | ···················· | ···················· | ···················· | ···················· |
| E | 5873 | hCoV-19/USA/WI-UW-802/2020\|EPI_ISL_495506\|2020-07- | ···················· | ···················· | ···················· | ···················· | ···················· | ···················· |
| E | 5874 | hCoV-19/USA/WI-UW-803/2020\|EPI_ISL_495507\|2020-07- | ···················· | ···················· | ···················· | ···················· | ···················· | ···················· |
| E | 5875 | hCoV-19/USA/IL-UW-804/2020\|EPI_ISL_495508\|2020-07- | ···················· | ···················· | ···················· | ···················· | ···················· | ···················· |
| E | 5876 | hCoV-19/USA/WI-UW-805/2020\|EPI_ISL_495509\|2020-07- | ···················· | ···················· | ···················· | ···················· | ···················· | ···················· |
| E | 5877 | hCoV-19/South Africa/KRISP-0344/2020\|EPI_ISL_49552 | ···················· | ···················· | ···················· | ···················· | ···················· | ···················· |
| E | 5879 | hCoV-19/South Africa/KRISP-0403/2020\|EPI_ISL_49552 | ···················· | ···················· | ···················· | ···················· | ···················· | ···················· |
| E | 5880 | hCoV-19/South Africa/KRISP-0404/2020\|EPI_ISL_49552 | ···················· | ···················· | ···················· | ···················· | ···················· | ···················· |
| E | 5881 | hCoV-19/South Africa/KRISP-0405/2020\|EPI_ISL_49553 | ···················· | ···················· | ···················· | ···················· | ···················· | ···················· |
| E | 5882 | hCoV-19/South Africa/KRISP-0409/2020\|EPI_ISL_49553 | ···················· | ···················· | ···················· | ···················· | ···················· | ···················· |
| E | 5883 | hCoV-19/South Africa/KRISP-0410/2020\|EPI_ISL_49553 | ···················· | ···················· | ···················· | ···················· | ···················· | ···················· |
| E | 5884 | hCoV-19/South Africa/KRISP-0411/2020\|EPI_ISL_49553 | ···················· | ···················· | ···················· | ···················· | ···················· | ···················· |
| E | 5885 | hCoV-19/South Africa/KRISP-0412/2020\|EPI_ISL_49553 | ···················· | ···················· | ···················· | ···················· | ···················· | ···················· |
| E | 5886 | hCoV-19/South Africa/KRISP-0413/2020\|EPI_ISL_49553 | ···················· | ···················· | ···················· | ···················· | ···················· | ···················· |
| E | 5887 | hCoV-19/South Africa/KRISP-0414/2020\|EPI_ISL_49553 | ···················· | ···················· | ···················· | ···················· | ···················· | ···················· |
| E | 5888 | hCoV-19/South Africa/KRISP-0415/2020\|EPI_ISL_49554 | ···················· | ···················· | ···················· | ···················· | ···················· | ···················· |
| E | 5889 | hCoV-19/USA/CA-ALSR-0522-SAN/2020\|EPI_ISL_483200\|2 | ···················· | ···················· | ···················· | ···················· | ···················· | ···················· |
| E | 5890 | hCoV-19/USA/CA-ALSR-0524-SAN/2020\|EPI_ISL_483201\|2 | ···················· | ···················· | ···················· | ···················· | ···················· | ···················· |
| E | 5891 | hCoV-19/USA/CA-ALSR-0525-IPL/2020\|EPI_ISL_483202\|2 | ···················· | ·A·················· | ···················· | ···················· | ···················· | ···················· |
| E | 5892 | hCoV-19/USA/CA-ALSR-0530-SAN/2020\|EPI_ISL_483204\|2 | ···················· | ···················· | ···················· | ···················· | ···················· | ···················· |
| E | 5893 | hCoV-19/USA/CA-ALSR-0531-SAN/2020\|EPI_ISL_483205\|2 | ···················· | ·A·················· | ···················· | ···················· | ···················· | ···················· |

| | | | | | | | | |
|---|---|---|---|---|---|---|---|---|
| E | 5894 hCoV-19/USA/CA-ALSR-0532-SAN/2020\|EPI_ISL_483206\|2 | ···················· | ···················· | ···················· | ···················· | ···················· | ···················· | |
| E | 5895 hCoV-19/USA/CA-ALSR-0534-IPL/2020\|EPI_ISL_483207\|2 | ···················· | ·A·················· | ···················· | ···················· | ···················· | ···················· | |
| E | 5896 hCoV-19/USA/CA-ALSR-0535-SAN/2020\|EPI_ISL_483208\|2 | ···················· | ·A·················· | ···················· | ···················· | ···················· | ···················· | |
| E | 5897 hCoV-19/USA/CA-ALSR-0538-SAN/2020\|EPI_ISL_483210\|2 | ···················· | ·A·················· | ···················· | ···················· | ···················· | ···················· | |
| E | 5898 hCoV-19/USA/CA-ALSR-0539-IPL/2020\|EPI_ISL_483211\|2 | ···················· | ·A·················· | ···················· | ···················· | ···················· | ···················· | |
| E | 5899 hCoV-19/USA/CA-ALSR-0541-SAN/2020\|EPI_ISL_483212\|2 | ···················· | ···················· | ···················· | ···················· | ···················· | ···················· | |
| E | 5900 hCoV-19/USA/CA-ALSR-0543-SAN/2020\|EPI_ISL_483213\|2 | ···················· | ·A·················· | ···················· | ···················· | ···················· | ···················· | |
| E | 5901 hCoV-19/USA/CA-ALSR-0550-SAN/2020\|EPI_ISL_483216\|2 | ···················· | ···················· | ···················· | ···················· | ···················· | ···················· | |
| E | 5902 hCoV-19/USA/CA-ALSR-0551-IPL/2020\|EPI_ISL_483217\|2 | ···················· | ···················· | ···················· | ···················· | ···················· | ···················· | |
| E | 5903 hCoV-19/USA/CA-ALSR-0555-SAN/2020\|EPI_ISL_483220\|2 | ···················· | ···················· | ···················· | ···················· | ···················· | ···················· | |
| E | 5904 hCoV-19/USA/CA-ALSR-0587-SAN/2020\|EPI_ISL_483243\|2 | ···················· | ···················· | ···················· | ···················· | ···················· | ···················· | |
| E | 5905 hCoV-19/USA/CA-ALSR-0588-IPL/2020\|EPI_ISL_483244\|2 | ···················· | ···················· | ···················· | ···················· | ···················· | ···················· | |
| E | 5906 hCoV-19/USA/CA-ALSR-0590-SAN/2020\|EPI_ISL_483245\|2 | ···················· | ···················· | ···················· | ···················· | ···················· | ···················· | |
| E | 5907 hCoV-19/USA/CA-ALSR-0593-SAN/2020\|EPI_ISL_483247\|2 | ···················· | ···················· | ···················· | ···················· | ···················· | ···················· | |
| E | 5908 hCoV-19/USA/CA-ALSR-0597-IPL/2020\|EPI_ISL_483248\|2 | ···················· | ·A·················· | ···················· | ···················· | ···················· | ···················· | |
| E | 5909 hCoV-19/USA/CA-ALSR-0600-IPL/2020\|EPI_ISL_483249\|2 | ···················· | ···················· | ···················· | ···················· | ···················· | ···················· | |
| E | 5910 hCoV-19/USA/CA-ALSR-0602-IPL/2020\|EPI_ISL_483250\|2 | ···················· | ·A·················· | ···················· | ···················· | ···················· | ···················· | |
| E | 5911 hCoV-19/USA/CA-ALSR-0607-IPL/2020\|EPI_ISL_483252\|2 | ···················· | ·A·················· | ···················· | ···················· | ···················· | ···················· | |
| E | 5912 hCoV-19/USA/CA-ALSR-0608-IPL/2020\|EPI_ISL_483253\|2 | ···················· | ·A·················· | ···················· | ···················· | ···················· | ···················· | |
| E | 5913 hCoV-19/USA/CA-ALSR-0609-SAN/2020\|EPI_ISL_483254\|2 | ···················· | ···················· | ···················· | ···················· | ···················· | ···················· | |
| E | 5914 hCoV-19/USA/CA-ALSR-0610-IPL/2020\|EPI_ISL_483255\|2 | ···················· | ···················· | ···················· | ···················· | ···················· | ···················· | |
| E | 5915 hCoV-19/USA/CA-ALSR-0611-SAN/2020\|EPI_ISL_483256\|2 | ···················· | ·A·················· | ···················· | ···················· | ···················· | ···················· | |
| E | 5916 hCoV-19/USA/CA-ALSR-0612-SAN/2020\|EPI_ISL_483257\|2 | ···················· | ···················· | ···················· | ···················· | ···················· | ···················· | |
| E | 5917 hCoV-19/USA/CA-ALSR-0614-SAN/2020\|EPI_ISL_483258\|2 | ···················· | ···················· | ···················· | ···················· | ···················· | ···················· | |
| E | 5918 hCoV-19/USA/CA-ALSR-0615-SAN/2020\|EPI_ISL_483259\|2 | ···················· | ···················· | ···················· | ···················· | ···················· | ···················· | |
| E | 5919 hCoV-19/USA/CA-ALSR-0616-SAN/2020\|EPI_ISL_483260\|2 | ···················· | ···················· | ···················· | ···················· | ···················· | ···················· | |
| E | 5920 hCoV-19/USA/CA-ALSR-0617-SAN/2020\|EPI_ISL_483261\|2 | ···················· | ···················· | ···················· | ···················· | ···················· | ···················· | |
| E | 5921 hCoV-19/USA/CA-ALSR-0621-IPL/2020\|EPI_ISL_483262\|2 | ···················· | ·A·················· | ···················· | ···················· | ···················· | ···················· | |
| E | 5922 hCoV-19/USA/CA-ALSR-0623-IPL/2020\|EPI_ISL_483263\|2 | ···················· | ···················· | ···················· | ···················· | ···················· | ···················· | |
| E | 5923 hCoV-19/USA/CA-ALSR-0624-IPL/2020\|EPI_ISL_483264\|2 | ···················· | ···················· | ···················· | ···················· | ···················· | ···················· | |
| E | 5924 hCoV-19/USA/CA-ALSR-0626-IPL/2020\|EPI_ISL_483265\|2 | ···················· | ·A·················· | ···················· | ···················· | ···················· | ···················· | |
| E | 5925 hCoV-19/USA/CA-ALSR-0627-IPL/2020\|EPI_ISL_483266\|2 | ···················· | ···················· | ···················· | ···················· | ···················· | ···················· | |
| E | 5926 hCoV-19/USA/CA-ALSR-0628-IPL/2020\|EPI_ISL_483267\|2 | ···················· | ···················· | ···················· | ···················· | ···················· | ···················· | |
| E | 5927 hCoV-19/USA/CA-ALSR-0629-IPL/2020\|EPI_ISL_483268\|2 | ···················· | ···················· | ···················· | ···················· | ···················· | ···················· | |
| E | 5928 hCoV-19/USA/CA-ALSR-0630-IPL/2020\|EPI_ISL_483269\|2 | ···················· | ···················· | ···················· | ···················· | ···················· | ···················· | |
| E | 5929 hCoV-19/USA/CA-ALSR-0631-IPL/2020\|EPI_ISL_483270\|2 | ···················· | ·A·················· | ···················· | ···················· | ···················· | ···················· | |
| E | 5930 hCoV-19/USA/CA-ALSR-0637-SAN/2020\|EPI_ISL_483272\|2 | ···················· | ·A·················· | ···················· | ···················· | ···················· | ···················· | |
| E | 5931 hCoV-19/USA/CA-ALSR-0638-IPL/2020\|EPI_ISL_483273\|2 | ···················· | ·A·················· | ···················· | ···················· | ···················· | ···················· | |
| E | 5932 hCoV-19/USA/CA-ALSR-0642-SAN/2020\|EPI_ISL_483275\|2 | ···················· | ···················· | ···················· | ···················· | ···················· | ···················· | |
| E | 5933 hCoV-19/USA/CA-ALSR-0643-IPL/2020\|EPI_ISL_483276\|2 | ···················· | ·A·················· | ···················· | ···················· | ···················· | ···················· | |
| E | 5934 hCoV-19/USA/CA-ALSR-0645-IPL/2020\|EPI_ISL_483277\|2 | ···················· | ·A·················· | ···················· | ···················· | ···················· | ···················· | |
| E | 5935 hCoV-19/USA/CA-ALSR-0649-SAN/2020\|EPI_ISL_483279\|2 | ···················· | ···················· | ···················· | ···················· | ···················· | ···················· | |
| E | 5936 hCoV-19/USA/CA-ALSR-0653-IPL/2020\|EPI_ISL_483281\|2 | ···················· | ·A·················· | ···················· | ···················· | ···················· | ···················· | |
| E | 5937 hCoV-19/USA/CA-ALSR-0657-IPL/2020\|EPI_ISL_483282\|2 | ···················· | ·A·················· | ···················· | ···················· | ···················· | ···················· | |
| E | 5938 hCoV-19/USA/CA-ALSR-0658-IPL/2020\|EPI_ISL_483283\|2 | ···················· | ···················· | ···················· | ···················· | ···················· | ···················· | |
| E | 5939 hCoV-19/USA/CA-ALSR-0659-IPL/2020\|EPI_ISL_483284\|2 | ···················· | ···················· | ···················· | ···················· | ···················· | ···················· | |
| E | 5940 hCoV-19/USA/CA-ALSR-0660-IPL/2020\|EPI_ISL_483285\|2 | ···················· | ·A·················· | ···················· | ···················· | ···················· | ···················· | |
| E | 5941 hCoV-19/USA/CA-ALSR-0661-IPL/2020\|EPI_ISL_483286\|2 | ···················· | ···················· | ···················· | ···················· | ···················· | ···················· | |
| E | 5942 hCoV-19/USA/CA-ALSR-0662-IPL/2020\|EPI_ISL_483287\|2 | ···················· | ·A·················· | ···················· | ···················· | ···················· | ···················· | |
| E | 5943 hCoV-19/USA/CA-ALSR-0663-IPL/2020\|EPI_ISL_483288\|2 | ···················· | ·A·················· | ···················· | ···················· | ···················· | ···················· | |
| E | 5944 hCoV-19/USA/CA-ALSR-0667-IPL/2020\|EPI_ISL_483289\|2 | ···················· | ···················· | ···················· | ···················· | ···················· | ···················· | |
| E | 5945 hCoV-19/USA/CA-ALSR-0668-SAN/2020\|EPI_ISL_483290\|2 | ···················· | ·A·················· | ···················· | ···················· | ···················· | ···················· | |
| E | 5946 hCoV-19/USA/CA-ALSR-0671-SAN/2020\|EPI_ISL_483291\|2 | ···················· | ···················· | ···················· | ···················· | ···················· | ···················· | |
| E | 5947 hCoV-19/USA/CA-ALSR-0673-SAN/2020\|EPI_ISL_483292\|2 | ···················· | ···················· | ···················· | ···················· | ···················· | ···················· | |
| E | 5948 hCoV-19/USA/CA-ALSR-0674-IPL/2020\|EPI_ISL_483293\|2 | ···················· | ·A·················· | ···················· | ···················· | ···················· | ···················· | |
| E | 5949 hCoV-19/USA/CA-ALSR-0675-IPL/2020\|EPI_ISL_483294\|2 | ···················· | ···················· | ···················· | ···················· | ···················· | ···················· | |
| E | 5950 hCoV-19/USA/CA-ALSR-0678-IPL/2020\|EPI_ISL_483295\|2 | ···················· | ·A·················· | ···················· | ···················· | ···················· | ···················· | |
| E | 5951 hCoV-19/USA/CA-ALSR-0679-IPL/2020\|EPI_ISL_483296\|2 | ···················· | ···················· | ···················· | ···················· | ···················· | ···················· | |
| E | 5952 hCoV-19/USA/CA-ALSR-0680-SAN/2020\|EPI_ISL_483297\|2 | ···················· | ···················· | ···················· | ···················· | ···················· | ···················· | |
| E | 5953 hCoV-19/USA/CA-ALSR-0681-IPL/2020\|EPI_ISL_483298\|2 | ···················· | ···················· | ···················· | ···················· | ···················· | ···················· | |
| E | 5954 hCoV-19/Scotland/CVR1947/2020\|EPI_ISL_484530\|2020- | ···················· | ···················A | ···················· | ···················· | ···················· | ···················· | 1 |
| E | 5955 hCoV-19/Scotland/CVR1948/2020\|EPI_ISL_484531\|2020- | ···················· | ···················· | ···················· | ···················· | ···················· | ···················· | |
| E | 5956 hCoV-19/Scotland/CVR1950/2020\|EPI_ISL_484532\|2020- | ···················· | ···················A | ···················· | ···················· | ···················· | ···················· | 1 |
| E | 5957 hCoV-19/Scotland/CVR1959/2020\|EPI_ISL_484533\|2020- | ···················· | ···················· | ···················· | ···················· | ···················· | ···················· | |
| E | 5958 hCoV-19/Scotland/CVR1967/2020\|EPI_ISL_484534\|2020- | ···················· | ···················· | ···················· | ···················· | ···················· | ···················· | |
| E | 5959 hCoV-19/Scotland/CVR1969/2020\|EPI_ISL_484535\|2020- | ···················· | ···················· | ···················· | ···················· | ···················· | ···················· | |
| E | 5960 hCoV-19/Scotland/CVR1974/2020\|EPI_ISL_484537\|2020- | ···················· | ···················· | ···················· | ···················· | ···················· | ···················· | |
| E | 5961 hCoV-19/Scotland/CVR1983/2020\|EPI_ISL_484538\|2020- | ···················· | ···················· | ···················· | ···················· | ···················· | ···················· | |
| E | 5962 hCoV-19/Scotland/CVR2021/2020\|EPI_ISL_484539\|2020- | ···················· | ···················· | ···················· | ···················· | ···················· | ···················· | |
| E | 5963 hCoV-19/Scotland/CVR2025/2020\|EPI_ISL_484540\|2020- | ···················· | ···················A | ···················· | ···················· | ···················· | ···················· | 1 |
| E | 5964 hCoV-19/Scotland/CVR2027/2020\|EPI_ISL_484541\|2020- | ···················· | ···················· | ···················· | ···················· | ···················· | ···················· | |
| E | 5965 hCoV-19/Scotland/CVR2028/2020\|EPI_ISL_484542\|2020- | ···················· | ···················A | ···················· | ···················· | ···················· | ···················· | 1 |
| E | 5966 hCoV-19/Scotland/CVR2039/2020\|EPI_ISL_484545\|2020- | ···················· | ···················· | ···················· | ···················· | ···················· | ···················· | |
| E | 5967 hCoV-19/Scotland/CVR2043/2020\|EPI_ISL_484546\|2020- | ···················· | ·A·················· | ···················· | ···················· | ···················· | ···················· | |
| E | 5968 hCoV-19/Scotland/CVR2045/2020\|EPI_ISL_484547\|2020- | ···················· | ·A·················· | ···················· | ···················· | ···················· | ···················· | |
| E | 5969 hCoV-19/Scotland/CVR2057/2020\|EPI_ISL_484548\|2020- | ···················· | ·A·················· | ···················· | ···················· | ···················· | ···················· | |
| E | 5970 hCoV-19/Scotland/CVR2065/2020\|EPI_ISL_484550\|2020- | ···················· | ···················· | ···················· | ···················· | ···················· | ···················· | |
| E | 5971 hCoV-19/Scotland/CVR2068/2020\|EPI_ISL_484551\|2020- | ···················· | ···················· | ···················· | ···················· | ···················· | ···················· | |
| E | 5972 hCoV-19/Scotland/CVR2298/2020\|EPI_ISL_484574\|2020- | ···················· | ···················· | ···················· | ···················· | ···················· | ···················· | |
| E | 5973 hCoV-19/Scotland/CVR2333/2020\|EPI_ISL_484576\|2020- | ···················· | ···················A | ···················· | ···················· | ···················· | ···················· | 1 |

| | | | | | | | | |
|---|---|---|---|---|---|---|---|---|
| E | 5974 hCoV-19/Scotland/CVR2334/2020\|EPI_ISL_484577\|2020- | .................... | .................... | ..................... | .................... | .................... | .................... | |
| E | 5975 hCoV-19/Scotland/CVR2373/2020\|EPI_ISL_484578\|2020- | .................... | .................... | ..................... | .................... | .................... | .................... | |
| E | 5976 hCoV-19/Scotland/CVR2374/2020\|EPI_ISL_484579\|2020- | .................... | .................... | ..................... | .................... | .................... | .................... | |
| E | 5977 hCoV-19/Scotland/CVR2377/2020\|EPI_ISL_484580\|2020- | .................... | .................... | ..................... | .................... | .................... | .................... | |
| E | 5978 hCoV-19/Scotland/CVR2381/2020\|EPI_ISL_484581\|2020- | .................... | .................... | ..................... | .................... | .................... | .................... | |
| E | 5979 hCoV-19/Scotland/CVR2389/2020\|EPI_ISL_484582\|2020- | .................... | .................... | ..................... | .................... | .................... | .................... | |
| E | 5980 hCoV-19/Scotland/CVR2402/2020\|EPI_ISL_484583\|2020- | .................... | .................... | ..................... | .................... | .................... | .................... | |
| E | 5981 hCoV-19/Scotland/CVR2418/2020\|EPI_ISL_484584\|2020- | .................... | .................... | ..................... | .................... | .................... | .................... | |
| E | 5982 hCoV-19/Scotland/CVR2425/2020\|EPI_ISL_484585\|2020- | .................... | .................... | ..................... | .................... | .................... | .................... | |
| E | 5983 hCoV-19/Scotland/CVR2426/2020\|EPI_ISL_484586\|2020- | .................... | .................... | ..................... | .................... | .................... | .................... | |
| E | 5984 hCoV-19/Scotland/CVR2434/2020\|EPI_ISL_484587\|2020- | .................... | .................... | ..................... | .................... | .................... | .................... | |
| E | 5985 hCoV-19/Scotland/CVR2435/2020\|EPI_ISL_484588\|2020- | .................... | .................... | ..................... | .................... | .................... | .................... | |
| E | 5986 hCoV-19/Scotland/CVR2449/2020\|EPI_ISL_484589\|2020- | .................... | ...................A | ..................... | .................... | .................... | .................... | 1 |
| E | 5987 hCoV-19/Scotland/CVR2465/2020\|EPI_ISL_484590\|2020- | .................... | .................... | ..................... | .................... | .................... | .................... | |
| E | 5988 hCoV-19/Scotland/CVR2481/2020\|EPI_ISL_484591\|2020- | .................... | .................... | ..................... | .................... | .................... | .................... | |
| E | 5989 hCoV-19/Scotland/CVR2493/2020\|EPI_ISL_484592\|2020- | .................... | .................... | ..................... | .................... | .................... | .................... | |
| E | 5990 hCoV-19/Scotland/CVR2500/2020\|EPI_ISL_484593\|2020- | .................... | .................... | ..................... | .................... | .................... | .................... | |
| E | 5991 hCoV-19/Scotland/CVR2516/2020\|EPI_ISL_484594\|2020- | .................... | .................... | ..................... | .................... | .................... | .................... | |
| E | 5992 hCoV-19/Scotland/CVR2519/2020\|EPI_ISL_484595\|2020- | .................... | .................... | ..................... | .................... | .................... | .................... | |
| E | 5993 hCoV-19/Scotland/CVR2525/2020\|EPI_ISL_484596\|2020- | .................... | .................... | ..................... | .................... | .................... | .................... | |
| E | 5994 hCoV-19/Scotland/CVR2530/2020\|EPI_ISL_484597\|2020- | .................... | .................... | ..................... | .................... | .................... | .................... | |
| E | 5995 hCoV-19/Scotland/CVR2535/2020\|EPI_ISL_484598\|2020- | .................... | ...................A | ..................... | .................... | .................... | .................... | 1 |
| E | 5996 hCoV-19/Scotland/CVR2540/2020\|EPI_ISL_484599\|2020- | .................... | ...................A | ..................... | .................... | .................... | .................... | 1 |
| E | 5997 hCoV-19/USA/VA-DCLS-0561/2020\|EPI_ISL_485861\|2020- | .................... | .................... | ..................... | .................... | .................... | .................... | |
| E | 5998 hCoV-19/USA/VA-DCLS-0563/2020\|EPI_ISL_485862\|2020- | .................... | .................... | ..................... | .................... | .................... | .................... | |
| E | 5999 hCoV-19/USA/VA-DCLS-0564/2020\|EPI_ISL_485863\|2020- | .................... | .................... | ..................... | .................... | .................... | .................... | |
| E | 6000 hCoV-19/USA/VA-DCLS-0565/2020\|EPI_ISL_485864\|2020- | .................... | .................... | ..................... | .................... | .................... | .................... | |
| E | 6001 hCoV-19/USA/VA-DCLS-0566/2020\|EPI_ISL_485865\|2020- | .................... | .................... | ..................... | .................... | .................... | .................... | |
| E | 6002 hCoV-19/USA/VA-DCLS-0568/2020\|EPI_ISL_485866\|2020- | .................... | .................... | ..................... | .................... | .................... | .................... | |
| E | 6003 hCoV-19/USA/VA-DCLS-0569/2020\|EPI_ISL_485867\|2020- | .................... | .................... | ..................... | .................... | .................... | .................... | |
| E | 6004 hCoV-19/USA/VA-DCLS-0570/2020\|EPI_ISL_485868\|2020- | .................... | .................... | ..................... | .................... | .................... | .................... | |
| E | 6005 hCoV-19/USA/VA-DCLS-0571/2020\|EPI_ISL_485869\|2020- | .................... | .................... | ..................... | .................... | .................... | .................... | |
| E | 6006 hCoV-19/USA/VA-DCLS-0572/2020\|EPI_ISL_485870\|2020- | .................... | .................... | ..................... | .................... | .................... | .................... | |
| E | 6007 hCoV-19/USA/VA-DCLS-0620/2020\|EPI_ISL_485871\|2020- | .................... | .................... | ..................... | .................... | .................... | .................... | |
| E | 6008 hCoV-19/USA/NM-UNM-00199/2020\|EPI_ISL_485872\|2020- | .................... | .................... | ..................... | .................... | .................... | .................... | |
| E | 6009 hCoV-19/USA/LA-GBCL-26492471/2020\|EPI_ISL_485874\|2 | .................... | .................... | ..................... | .................... | .................... | .................... | |
| E | 6010 hCoV-19/USA/LA-GBCL-26492486/2020\|EPI_ISL_485876\|2 | .................... | .................... | ..................... | .................... | .................... | .................... | |
| E | 6011 hCoV-19/USA/LA-GBCL-26512986/2020\|EPI_ISL_485879\|2 | .................... | .................... | ..................... | .................... | .................... | .................... | |
| E | 6012 hCoV-19/USA/LA-GBCL-26513051/2020\|EPI_ISL_485881\|2 | .................... | .................... | ..................... | .................... | .................... | .................... | |
| E | 6013 hCoV-19/England/NORT-29E83A/2020\|EPI_ISL_488127\|20 | .................... | ...................A | ..................... | .................... | .................... | .................... | 1 |
| E | 6014 hCoV-19/England/NORT-296E5E/2020\|EPI_ISL_488128\|20 | .................... | .................... | ..................... | .................... | .................... | .................... | |
| E | 6015 hCoV-19/England/NORT-297B83/2020\|EPI_ISL_488129\|20 | .................... | .................... | ..................... | .................... | .................... | .................... | |
| E | 6016 hCoV-19/England/NORT-297048/2020\|EPI_ISL_488130\|20 | .................... | .................... | ..................... | .................... | .................... | .................... | |
| E | 6017 hCoV-19/England/NORT-29D9A1/2020\|EPI_ISL_488131\|20 | .................... | .................... | ..................... | .................... | .................... | .................... | |
| E | 6018 hCoV-19/England/NORT-29C84B/2020\|EPI_ISL_488132\|20 | .................... | .................... | ..................... | .................... | .................... | .................... | |
| E | 6019 hCoV-19/England/NORT-29DD96/2020\|EPI_ISL_488133\|20 | .................... | .................... | ..................... | .................... | .................... | .................... | |
| E | 6020 hCoV-19/England/NORT-29700C/2020\|EPI_ISL_488136\|20 | .................... | .................... | ..................... | .................... | .................... | .................... | |
| E | 6021 hCoV-19/England/NORT-296DAC/2020\|EPI_ISL_488137\|20 | .................... | .................... | ..................... | .................... | .................... | .................... | |
| E | 6022 hCoV-19/Scotland/GCVR-171A74/2020\|EPI_ISL_489401\|2 | .................... | .A.................. | ..................... | .................... | .................... | .................... | |
| E | 6023 hCoV-19/Scotland/GCVR-171A38/2020\|EPI_ISL_489407\|2 | .................... | ...................A | ..................... | .................... | .................... | .................... | 1 |
| E | 6024 hCoV-19/Scotland/GCVR-171AFC/2020\|EPI_ISL_489408\|2 | .................... | .................... | ..................... | .................... | .................... | .................... | |
| E | 6025 hCoV-19/England/CAMB-1AD5C9/2020\|EPI_ISL_489409\|20 | .................... | .................... | ..................... | .................... | .................... | .................... | |
| E | 6026 hCoV-19/England/CAMB-1AC6A9/2020\|EPI_ISL_489411\|20 | .................... | .................... | ..................... | .................... | .................... | .................... | |
| E | 6027 hCoV-19/England/CAMB-1AC9BF/2020\|EPI_ISL_489414\|20 | .................... | .................... | ..................... | .................... | .................... | .................... | |
| E | 6028 hCoV-19/England/CAMB-1AC4CD/2020\|EPI_ISL_489415\|20 | .................... | .................... | ..................... | .................... | .................... | .................... | |
| E | 6029 hCoV-19/England/CAMB-1AD550/2020\|EPI_ISL_489416\|20 | .................... | .................... | ..................... | .................... | .................... | .................... | |
| E | 6030 hCoV-19/Scotland/GCVR-17193B/2020\|EPI_ISL_489418\|2 | .................... | .................... | ..................... | .................... | .................... | .................... | |
| E | 6031 hCoV-19/England/CAMB-1AC700/2020\|EPI_ISL_489420\|20 | .................... | .................... | ..................... | .................... | .................... | .................... | |
| E | 6032 hCoV-19/England/CAMB-1AC982/2020\|EPI_ISL_489458\|20 | .................... | .................... | ..................... | .................... | .................... | .................... | |
| E | 6033 hCoV-19/England/CAMB-1ACD77/2020\|EPI_ISL_489461\|20 | .................... | .................... | ..................... | .................... | .................... | .................... | |
| E | 6034 hCoV-19/England/CAMB-1AD5BA/2020\|EPI_ISL_489462\|20 | .................... | .................... | ..................... | .................... | .................... | .................... | |
| E | 6035 hCoV-19/England/CAMB-1AC427/2020\|EPI_ISL_489465\|20 | .................... | .................... | ..................... | .................... | .................... | .................... | |
| E | 6036 hCoV-19/England/CAMB-1ACB8C/2020\|EPI_ISL_489466\|20 | .................... | .................... | ..................... | .................... | .................... | .................... | |
| E | 6037 hCoV-19/England/LIVE-A41DE/2020\|EPI_ISL_490408\|202 | .................... | .................... | ..................... | .................... | .................... | .................... | |
| E | 6038 hCoV-19/England/LIVE-A41ED/2020\|EPI_ISL_490409\|202 | .................... | .................... | ..................... | .................... | .................... | .................... | |
| E | 6039 hCoV-19/England/LOND-124C829/2020\|EPI_ISL_490436\|2 | .................... | .................... | ..................... | .................... | .................... | .................... | |
| E | 6040 hCoV-19/England/LOND-124C874/2020\|EPI_ISL_490438\|2 | .................... | .................... | ..................... | .................... | .................... | .................... | |
| E | 6041 hCoV-19/England/LOND-124C917/2020\|EPI_ISL_490439\|2 | .................... | .............A...... | ..................... | .................... | .................... | .................... | |
| E | 6042 hCoV-19/Scotland/EDB6439/2020\|EPI_ISL_491700\|2020- | .................... | .................... | ..................... | .................... | .................... | .................... | |
| E | 6043 hCoV-19/England/202881370/2020\|EPI_ISL_491709\|2020 | .................... | .................... | ..................... | .................... | .................... | .................... | |
| E | 6044 hCoV-19/England/202881376/2020\|EPI_ISL_491712\|2020 | .................... | .................... | ..................... | .................... | .................... | .................... | |
| E | 6045 hCoV-19/England/202881378/2020\|EPI_ISL_491713\|2020 | .................... | .................... | ..................... | .................... | .................... | .................... | |
| E | 6046 hCoV-19/England/202881379/2020\|EPI_ISL_491714\|2020 | .................... | .................... | ..................... | .................... | .................... | .................... | |
| E | 6047 hCoV-19/England/202881380/2020\|EPI_ISL_491715\|2020 | .................... | .................... | ..................... | .................... | .................... | .................... | |
| E | 6048 hCoV-19/England/202881382/2020\|EPI_ISL_491716\|2020 | .................... | .................... | ..................... | .................... | .................... | .................... | |
| E | 6049 hCoV-19/England/202881383/2020\|EPI_ISL_491717\|2020 | .................... | .................... | ..................... | .................... | .................... | .................... | |
| E | 6050 hCoV-19/England/202881384/2020\|EPI_ISL_491718\|2020 | .................... | .................... | ..................... | .................... | .................... | .................... | |
| E | 6051 hCoV-19/England/202881385/2020\|EPI_ISL_491719\|2020 | .................... | .................... | ..................... | .................... | .................... | .................... | |
| E | 6052 hCoV-19/England/202881386/2020\|EPI_ISL_491720\|2020 | .................... | .................... | ..................... | .................... | .................... | .................... | |
| E | 6053 hCoV-19/England/202881387/2020\|EPI_ISL_491721\|2020 | .................... | .................... | ..................... | .................... | .................... | .................... | |

E 6054 hCoV-19/England/202881388/2020|EPI_ISL_491722|2020 ···················· ···················· ····················· ···················· ···················· ····················
E 6055 hCoV-19/England/202881389/2020|EPI_ISL_491723|2020 ···················· ···················· ····················· ···················· ···················· ····················
E 6056 hCoV-19/England/202881390/2020|EPI_ISL_491724|2020 ···················· ···················· ····················· ···················· ···················· ····················
E 6057 hCoV-19/England/202881391/2020|EPI_ISL_491725|2020 ···················· ···················· ····················· ···················· ···················· ····················
E 6058 hCoV-19/England/202881392/2020|EPI_ISL_491726|2020 ···················· ···················· ····················· ···················· ···················· ····················
E 6059 hCoV-19/England/202881393/2020|EPI_ISL_491727|2020 ···················· ···················· ····················· ···················· ···················· ····················
E 6060 hCoV-19/England/202881394/2020|EPI_ISL_491728|2020 ···················· ···················· ····················· ···················· ···················· ····················
E 6061 hCoV-19/England/202881395/2020|EPI_ISL_491729|2020 ···················· ···················· ····················· ···················· ···················· ····················
E 6062 hCoV-19/England/202881397/2020|EPI_ISL_491731|2020 ···················· ···················· ····················· ···················· ···················· ····················
E 6063 hCoV-19/England/202881398/2020|EPI_ISL_491732|2020 ···················· ···················· ····················· ···················· ···················· ····················
E 6064 hCoV-19/England/202881399/2020|EPI_ISL_491733|2020 ···················· ···················· ····················· ···················· ···················· ····················
E 6065 hCoV-19/England/202881400/2020|EPI_ISL_491734|2020 ···················· ···················· ····················· ···················· ···················· ····················
E 6066 hCoV-19/England/202881401/2020|EPI_ISL_491735|2020 ···················· ···················· ····················· ···················· ···················· ····················
E 6067 hCoV-19/India/TG-CCMB-M664/2020|EPI_ISL_495222|202 ···················· ···················· ····················· ···················· ···················· ····················
E 6068 hCoV-19/India/TG-CCMB-M686/2020|EPI_ISL_495223|202 ···················· ···················· ····················· ···················· ···················· ····················
E 6069 hCoV-19/India/TG-CCMB-M696/2020|EPI_ISL_495224|202 ···················· ···················· ····················· ···················· ···················· ····················
E 6072 hCoV-19/India/TG-CCMB-M70/2020|EPI_ISL_495227|2020 ···················· ·A·················· ····················· ···················· ···················· ··A·················
E 6073 hCoV-19/India/TG-CCMB-M700/2020|EPI_ISL_495228|202 ···················· ···················· ····················· ···················· ···················· ····················
E 6074 hCoV-19/India/TG-CCMB-M711/2020|EPI_ISL_495229|202 ···················· ···················· ····················· ···················· ···················· ····················
E 6075 hCoV-19/India/TG-CCMB-M712/2020|EPI_ISL_495230|202 ···················· ···················· ····················· ···················· ···················· ····················
E 6076 hCoV-19/India/TG-CCMB-M717/2020|EPI_ISL_495231|202 ···················· ···················· ····················· ···················· ···················· ····················
E 6077 hCoV-19/India/TG-CCMB-M722/2020|EPI_ISL_495232|202 ···················· ···················· ····················· ···················· ···················· ····················
E 6078 hCoV-19/India/TG-CCMB-M733/2020|EPI_ISL_495233|202 ···················· ···················· ····················· ···················· ···················· ····················
E 6079 hCoV-19/India/TG-CCMB-M739/2020|EPI_ISL_495234|202 ···················· ···················· ····················· ···················· ···················· ····················
E 6080 hCoV-19/India/TG-CCMB-M740/2020|EPI_ISL_495235|202 ···················· ···················· ····················· ···················· ···················· ····················
E 6081 hCoV-19/India/TG-CCMB-M750/2020|EPI_ISL_495236|202 ···················· ···················· ····················· ···················· ···················· ····················
E 6082 hCoV-19/India/TG-CCMB-M762/2020|EPI_ISL_495237|202 ···················· ···················· ····················· ···················· ···················· ····················
E 6083 hCoV-19/India/TG-CCMB-M772/2020|EPI_ISL_495238|202 ···················· ···················· ····················· ···················· ···················· ····················
E 6084 hCoV-19/India/TG-CCMB-M779/2020|EPI_ISL_495239|202 ···················· ·A·················· ····················· ···················· ···················· ··A·················
E 6085 hCoV-19/India/TG-CCMB-M780/2020|EPI_ISL_495240|202 ···················· ···················· ····················· ···················· ···················· ····················
E 6086 hCoV-19/India/TG-CCMB-M785/2020|EPI_ISL_495241|202 ···················· ···················· ····················· ···················· ···················· ····················
E 6087 hCoV-19/India/TG-CCMB-M79/2020|EPI_ISL_495242|2020 ···················· ···················· ····················· ···················· ···················· ····················
E 6088 hCoV-19/India/TG-CCMB-M80/2020|EPI_ISL_495243|2020 ···················· ···················· ····················· ···················· ···················· ····················
E 6089 hCoV-19/India/TG-CCMB-M804/2020|EPI_ISL_495244|202 ···················· ·A·················· ····················· ···················· ···················· ··A·················
E 6090 hCoV-19/India/TG-CCMB-M806/2020|EPI_ISL_495245|202 ···················· ···················· ····················· ···················· ···················· ····················
E 6091 hCoV-19/India/TG-CCMB-M81/2020|EPI_ISL_495246|2020 ···················· ···················· ····················· ···················· ···················· ····················
E 6092 hCoV-19/India/TG-CCMB-M812/2020|EPI_ISL_495247|202 ···················· ···················· ····················· ···················· ···················· ····················
E 6093 hCoV-19/India/TG-CCMB-M817/2020|EPI_ISL_495248|202 ···················· ···················· ····················· ···················· ···················· ····················
E 6094 hCoV-19/India/TG-CCMB-M818/2020|EPI_ISL_495249|202 ···················· ···················· ····················· ···················· ···················· ····················
E 6095 hCoV-19/India/TG-CCMB-M82/2020|EPI_ISL_495250|2020 ···················· ···················· ····················· ···················· ···················· ····················
E 6096 hCoV-19/India/TG-CCMB-M830/2020|EPI_ISL_495251|202 ···················· ···················· ····················· ···················· ···················· ····················
E 6097 hCoV-19/India/TG-CCMB-M838/2020|EPI_ISL_495252|202 ···················· ···················· ····················· ···················· ···················· ····················
E 6098 hCoV-19/India/TG-CCMB-M840/2020|EPI_ISL_495253|202 ···················· ···················· ····················· ···················· ···················· ····················
E 6099 hCoV-19/India/TG-CCMB-M841/2020|EPI_ISL_495254|202 ···················· ···················· ····················· ···················· ···················· ····················
E 6100 hCoV-19/India/TG-CCMB-M842/2020|EPI_ISL_495255|202 ···················· ···················· ····················· ···················· ···················· ····················
E 6101 hCoV-19/India/TG-CCMB-M844/2020|EPI_ISL_495256|202 ···················· ···················· ····················· ···················· ···················· ····················
E 6102 hCoV-19/India/TG-CCMB-M845/2020|EPI_ISL_495257|202 ···················· ···················· ····················· ···················· ···················· ····················
E 6103 hCoV-19/India/TG-CCMB-M860/2020|EPI_ISL_495258|202 ···················· ···················· ····················· ···················· ···················· ····················
E 6104 hCoV-19/India/TG-CCMB-M882/2020|EPI_ISL_495259|202 ···················· ···················· ····················· ···················· ···················· ····················
E 6105 hCoV-19/India/TG-CCMB-M891/2020|EPI_ISL_495260|202 ···················· ···················· ····················· ···················· ···················· ····················
E 6106 hCoV-19/India/TG-CCMB-M892/2020|EPI_ISL_495261|202 ···················· ···················· ····················· ···················· ···················· ····················
E 6107 hCoV-19/India/TG-CCMB-M900/2020|EPI_ISL_495262|202 ···················· ···················· ····················· ···················· ···················· ····················
E 6108 hCoV-19/India/TG-CCMB-M925/2020|EPI_ISL_495263|202 ···················· ···················· ····················· ···················· ···················· ····················
E 6109 hCoV-19/India/TG-CCMB-M930/2020|EPI_ISL_495264|202 ···················· ···················· ····················· ···················· ···················· ····················
E 6110 hCoV-19/India/TG-CCMB-M931/2020|EPI_ISL_495265|202 ···················· ···················· ····················· ···················· ···················· ····················
E 6111 hCoV-19/India/TG-CCMB-M950/2020|EPI_ISL_495266|202 ···················· ···················· ····················· ···················· ···················· ··A·················
E 6112 hCoV-19/India/TG-CCMB-M969/2020|EPI_ISL_495267|202 ···················· ···················· ····················· ···················· ···················· ····················
E 6113 hCoV-19/India/TG-CCMB-M971/2020|EPI_ISL_495268|202 ···················· ···················· ····················· ···················· ···················· ····················
E 6114 hCoV-19/India/TG-CCMB-M980/2020|EPI_ISL_495269|202 ···················· ···················· ····················· ···················· ···················· ····················
E 6115 hCoV-19/India/TG-CCMB-M995/2020|EPI_ISL_495270|202 ···················· ···················· ····················· ···················· ···················· ····················
E 6116 hCoV-19/India/TG-CCMB-N18/2020|EPI_ISL_495271|2020 ···················· ···················· ····················· ···················· ···················· ····················
E 6117 hCoV-19/India/TG-CCMB-N4/2020|EPI_ISL_495272|2020- ···················· ···················· ····················· ···················· ···················· ····················
E 6118 hCoV-19/India/TG-CCMB-N7/2020|EPI_ISL_495273|2020- ···················· ···················· ····················· ···················· ···················· ····················
E 6119 hCoV-19/India/TG-CCMB-OM1/2020|EPI_ISL_495274|2020 ···················· ···················· ····················· ···················· ···················· ····················
E 6120 hCoV-19/India/TG-CCMB-OM1-ONT/2020|EPI_ISL_495275| ···················· ···················· ····················· ···················· ···················· ····················
E 6121 hCoV-19/India/TG-CCMB-OM10/2020|EPI_ISL_495276|202 ···················· ···················· ····················· ···················· ···················· ····················
E 6122 hCoV-19/India/TG-CCMB-OM11/2020|EPI_ISL_495277|202 ···················· ···················· ····················· ···················· ···················· ····················
E 6123 hCoV-19/India/TG-CCMB-OM12/2020|EPI_ISL_495278|202 ···················· ···················· ····················· ···················· ···················· ····················
E 6124 hCoV-19/India/TG-CCMB-OM13/2020|EPI_ISL_495279|202 ···················· ···················· ····················· ···················· ···················· ····················
E 6125 hCoV-19/India/TG-CCMB-OM14/2020|EPI_ISL_495280|202 ···················· ···················· ····················· ···················· ···················· ····················
E 6126 hCoV-19/India/TG-CCMB-OM15/2020|EPI_ISL_495281|202 ···················· ···················· ····················· ···················· ···················· ····················
E 6127 hCoV-19/India/TG-CCMB-OM16/2020|EPI_ISL_495282|202 ···················· ···················· ····················· ···················· ···················· ····················
E 6128 hCoV-19/India/TG-CCMB-OM17/2020|EPI_ISL_495283|202 ···················· ···················· ····················· ···················· ···················· ····················
E 6129 hCoV-19/India/TG-CCMB-OM18/2020|EPI_ISL_495284|202 ···················· ···················· ····················· ···················· ···················· ····················
E 6130 hCoV-19/India/TG-CCMB-OM19/2020|EPI_ISL_495285|202 ···················· ···················· ····················· ···················· ···················· ····················
E 6131 hCoV-19/India/TG-CCMB-OM2/2020|EPI_ISL_495286|2020 ···················· ···················· ····················· ···················· ···················· ····················
E 6132 hCoV-19/India/TG-CCMB-OM2-ONT/2020|EPI_ISL_495287| ···················· ···················· ····················· ···················· ···················· ····················
E 6133 hCoV-19/India/TG-CCMB-OM20/2020|EPI_ISL_495288|202 ···················· ···················· ····················· ···················· ···················· ····················
E 6134 hCoV-19/India/TG-CCMB-OM3/2020|EPI_ISL_495289|2020 ···················· ···················· ····················· ···················· ···················· ····················
E 6135 hCoV-19/India/TG-CCMB-OM4/2020|EPI_ISL_495290|2020 ···················· ···················· ····················· ···················· ···················· ····················

| | | | | | | | | | |
|---|---|---|---|---|---|---|---|---|---|
| E | 6136 | hCoV-19/India/TG-CCMB-OM4-ONT/2020\|EPI_ISL_495291\| | ···················· | ···················· | ···················· | ···················· | ···················· | ···················· | |
| E | 6137 | hCoV-19/India/TG-CCMB-OM5/2020\|EPI_ISL_495292\|2020 | ···················· | ···················· | ···················· | ···················· | ···················· | ···················· | |
| E | 6138 | hCoV-19/India/TG-CCMB-OM6/2020\|EPI_ISL_495293\|2020 | ···················· | ···················· | ···················· | ···················· | ···················· | ···················· | |
| E | 6139 | hCoV-19/India/TG-CCMB-OM6-ONT/2020\|EPI_ISL_495294\| | ···················· | ···················· | ···················· | ···················· | ···················· | ···················· | |
| E | 6140 | hCoV-19/India/TG-CCMB-OM7/2020\|EPI_ISL_495295\|2020 | ···················· | ···················· | ···················· | ···················· | ···················· | ···················· | |
| E | 6141 | hCoV-19/India/TG-CCMB-OM8/2020\|EPI_ISL_495296\|2020 | ···················· | ···················· | ···················· | ···················· | ···················· | ···················· | |
| E | 6142 | hCoV-19/India/TG-CCMB-OM9/2020\|EPI_ISL_495297\|2020 | ···················· | ···················· | ···················· | ···················· | ···················· | ···················· | |
| E | 6143 | hCoV-19/USA/VA-DCLS-0625/2020\|EPI_ISL_495298\|2020- | ···················· | ···················· | ···················· | ···················· | ···················· | ···················· | |
| E | 6144 | hCoV-19/USA/VA-DCLS-0626/2020\|EPI_ISL_495299\|2020- | ···················· | ···················· | ···················· | ···················· | ···················· | ···················· | |
| E | 6145 | hCoV-19/USA/CA-ALSR-0547-IPL/2020\|EPI_ISL_483214\|2 | ···················· | ···················· | ···················· | ···················· | ···················· | ···················· | |
| E | 6146 | hCoV-19/USA/CA-ALSR-0548-IPL/2020\|EPI_ISL_483215\|2 | ···················· | ·A·················· | ···················· | ···················· | ···················· | ···················· | |
| E | 6147 | hCoV-19/USA/CA-ALSR-0552-IPL/2020\|EPI_ISL_483218\|2 | ···················· | ·A·················· | ···················· | ···················· | ···················· | ···················· | |
| E | 6148 | hCoV-19/USA/CA-ALSR-0553-IPL/2020\|EPI_ISL_483219\|2 | ···················· | ·A·················· | ···················· | ···················· | ···················· | ···················· | |
| E | 6149 | hCoV-19/USA/CA-ALSR-0557-SAN/2020\|EPI_ISL_483221\|2 | ···················· | ·A·················· | ···················· | ···················· | ···················· | ···················· | |
| E | 6150 | hCoV-19/USA/CA-ALSR-0558-IPL/2020\|EPI_ISL_483222\|2 | ···················· | ·A·················· | ···················· | ···················· | ···················· | ···················· | |
| E | 6151 | hCoV-19/USA/CA-ALSR-0562-SAN/2020\|EPI_ISL_483224\|2 | ···················· | ·A·················· | ···················· | ···················· | ···················· | ···················· | |
| E | 6152 | hCoV-19/USA/CA-ALSR-0563-SAN/2020\|EPI_ISL_483225\|2 | ···················· | ·A·················· | ···················· | ···················· | ···················· | ···················· | |
| E | 6153 | hCoV-19/USA/CA-ALSR-0565-SAN/2020\|EPI_ISL_483226\|2 | ···················· | ···················· | ···················· | ···················· | ···················· | ···················· | |
| E | 6154 | hCoV-19/USA/CA-ALSR-0566-IPL/2020\|EPI_ISL_483227\|2 | ···················· | ·A·················· | ···················· | ···················· | ···················· | ···················· | |
| E | 6155 | hCoV-19/USA/CA-ALSR-0567-IPL/2020\|EPI_ISL_483228\|2 | ···················· | ·A·················· | ···················· | ···················· | ···················· | ···················· | |
| E | 6156 | hCoV-19/USA/CA-ALSR-0568-SAN/2020\|EPI_ISL_483229\|2 | ···················· | ·A·················· | ···················· | ···················· | ···················· | ···················· | |
| E | 6157 | hCoV-19/USA/CA-ALSR-0570-IPL/2020\|EPI_ISL_483230\|2 | ···················· | ·A·················· | ···················· | ···················· | ···················· | ···················· | |
| E | 6158 | hCoV-19/USA/CA-ALSR-0571-IPL/2020\|EPI_ISL_483231\|2 | ···················· | ·A·················· | ···················· | ···················· | ···················· | ···················· | |
| E | 6159 | hCoV-19/USA/CA-ALSR-0576-SAN/2020\|EPI_ISL_483232\|2 | ···················· | ·A·················· | ···················· | ···················· | ···················· | ···················· | |
| E | 6160 | hCoV-19/USA/CA-ALSR-0577-IPL/2020\|EPI_ISL_483233\|2 | ···················· | ·A·················· | ···················· | ···················· | ···················· | ···················· | |
| E | 6161 | hCoV-19/USA/CA-ALSR-0578-SAN/2020\|EPI_ISL_483234\|2 | ···················· | ···················· | ···················· | ···················· | ···················· | ···················· | |
| E | 6162 | hCoV-19/USA/CA-ALSR-0579-SAN/2020\|EPI_ISL_483235\|2 | ···················· | ·A·················· | ···················· | ···················· | ···················· | ···················· | |
| E | 6163 | hCoV-19/USA/CA-ALSR-0580-IPL/2020\|EPI_ISL_483236\|2 | ···················· | ·A·················· | ···················· | ···················· | ···················· | ···················· | |
| E | 6164 | hCoV-19/USA/CA-ALSR-0581-IPL/2020\|EPI_ISL_483237\|2 | ···················· | ·A·················· | ···················· | ···················· | ···················· | ···················· | |
| E | 6165 | hCoV-19/USA/CA-ALSR-0582-SAN/2020\|EPI_ISL_483238\|2 | ···················· | ·A·················· | ···················· | ···················· | ···················· | ···················· | |
| E | 6166 | hCoV-19/USA/CA-ALSR-0583-SAN/2020\|EPI_ISL_483239\|2 | ···················· | ···················· | ···················· | ···················· | ···················· | ···················· | |
| E | 6167 | hCoV-19/USA/CA-ALSR-0584-SAN/2020\|EPI_ISL_483240\|2 | ···················· | ···················· | ···················· | ···················· | ···················· | ···················· | |
| E | 6168 | hCoV-19/USA/CA-ALSR-0585-SAN/2020\|EPI_ISL_483241\|2 | ···················· | ·A·················· | ···················· | ···················· | ···················· | ···················· | |
| E | 6169 | hCoV-19/USA/CA-ALSR-0586-SAN/2020\|EPI_ISL_483242\|2 | ···················· | ···················· | ···················· | ···················· | ···················· | ···················· | |
| E | 6170 | hCoV-19/England/SHEF-CDC47/2020\|EPI_ISL_484505\|202 | ···················· | ···················· | ···················· | ···················· | ···················· | ···················· | |
| E | 6171 | hCoV-19/England/SHEF-CFD7F/2020\|EPI_ISL_484506\|202 | ···················· | ···················· | ···················· | ···················· | ···················· | ···················· | |
| E | 6172 | hCoV-19/England/SHEF-CFFA6/2020\|EPI_ISL_484508\|202 | ···················· | ···················· | ···················· | ···················· | ···················· | ···················· | |
| E | 6173 | hCoV-19/England/SHEF-D0458/2020\|EPI_ISL_484511\|202 | ···················· | ···················· | ···················· | ···················· | ···················· | ···················· | |
| E | 6174 | hCoV-19/England/SHEF-D125D/2020\|EPI_ISL_484514\|202 | ···················· | ···················· | ···················· | ···················· | ···················· | ···················· | |
| E | 6175 | hCoV-19/England/SHEF-D1466/2020\|EPI_ISL_484515\|202 | ···················· | ···················· | ···················· | ···················· | ···················· | ···················· | |
| E | 6176 | hCoV-19/England/SHEF-D1AEC/2020\|EPI_ISL_484516\|202 | ···················· | ···················· | ···················· | ···················· | ···················· | ···················· | |
| E | 6177 | hCoV-19/Scotland/CVR1872/2020\|EPI_ISL_484519\|2020- | ···················· | ···················· | ···················· | ···················· | ···················· | ···················· | |
| E | 6178 | hCoV-19/Scotland/CVR1874/2020\|EPI_ISL_484520\|2020- | ···················· | ···················· | ···················· | ···················· | ···················· | ···················· | |
| E | 6179 | hCoV-19/Scotland/CVR1884/2020\|EPI_ISL_484521\|2020- | ···················· | ···················· | ···················· | ···················· | ···················· | ···················· | |
| E | 6181 | hCoV-19/Scotland/CVR1890/2020\|EPI_ISL_484523\|2020- | ···················· | ···················· | ···················· | ···················· | ···················· | ···················· | |
| E | 6182 | hCoV-19/Scotland/CVR1928/2020\|EPI_ISL_484524\|2020- | ···················· | ···················· | ···················· | ···················· | ···················· | ···················· | |
| E | 6183 | hCoV-19/Scotland/CVR1932/2020\|EPI_ISL_484525\|2020- | ···················· | ···················· | ···················· | ···················· | ···················· | ···················· | |
| E | 6184 | hCoV-19/Scotland/CVR1933/2020\|EPI_ISL_484526\|2020- | ···················· | ···················· | ···················· | ···················· | ···················· | ···················· | |
| E | 6185 | hCoV-19/Scotland/CVR1939/2020\|EPI_ISL_484527\|2020- | ···················· | ···················· | ···················· | ···················· | ···················· | ···················· | |
| E | 6186 | hCoV-19/Scotland/CVR1941/2020\|EPI_ISL_484528\|2020- | ···················· | ·A·················· | ···················· | ···················· | ···················· | ···················· | |
| E | 6187 | hCoV-19/Scotland/CVR1945/2020\|EPI_ISL_484529\|2020- | ···················· | ·A·················· | ···················· | ···················· | ···················· | ···················· | |
| E | 6188 | hCoV-19/Scotland/CVR2030/2020\|EPI_ISL_484543\|2020- | ···················· | ···················· | ···················· | ···················· | ···················· | ···················· | |
| E | 6189 | hCoV-19/Scotland/CVR2035/2020\|EPI_ISL_484544\|2020- | ···················· | ·A·················· | ···················· | ···················· | ···················· | ···················· | |
| E | 6190 | hCoV-19/Scotland/CVR2062/2020\|EPI_ISL_484549\|2020- | ···················· | ·A·················· | ···················· | ···················· | ···················· | ···················· | |
| E | 6191 | hCoV-19/Scotland/CVR2072/2020\|EPI_ISL_484552\|2020- | ···················· | ·A·················· | ···················· | ···················· | ···················· | ···················· | |
| E | 6192 | hCoV-19/Scotland/CVR2079/2020\|EPI_ISL_484554\|2020- | ···················· | ···················· | ···················· | ···················· | ···················· | ···················· | |
| E | 6193 | hCoV-19/Scotland/CVR2080/2020\|EPI_ISL_484555\|2020- | ···················· | ···················A | ···················· | ···················· | ···················· | ···················· | 1 |
| E | 6194 | hCoV-19/Scotland/CVR2082/2020\|EPI_ISL_484556\|2020- | ···················· | ···················· | ···················· | ···················· | ···················· | ···················· | |
| E | 6195 | hCoV-19/Scotland/CVR2083/2020\|EPI_ISL_484557\|2020- | ···················· | ···················· | ···················· | ···················· | ···················· | ···················· | |
| E | 6196 | hCoV-19/Scotland/CVR2084/2020\|EPI_ISL_484558\|2020- | ···················· | ···················· | ···················· | ···················· | ···················· | ···················· | |
| E | 6197 | hCoV-19/Scotland/CVR2103/2020\|EPI_ISL_484559\|2020- | ···················· | ···················· | ···················· | ···················· | ···················· | ···················· | |
| E | 6198 | hCoV-19/Scotland/CVR2107/2020\|EPI_ISL_484560\|2020- | ···················· | ···················· | ···················· | ···················· | ···················· | ···················· | |
| E | 6199 | hCoV-19/Scotland/CVR2113/2020\|EPI_ISL_484561\|2020- | ···················· | ···················A | ···················· | ···················· | ···················· | ···················· | 1 |
| E | 6200 | hCoV-19/Scotland/CVR2115/2020\|EPI_ISL_484562\|2020- | ···················· | ·A·················· | ···················· | ···················· | ···················· | ···················· | |
| E | 6201 | hCoV-19/Scotland/CVR2147/2020\|EPI_ISL_484563\|2020- | ···················· | ···················· | ···················· | ···················· | ···················· | ···················· | |
| E | 6202 | hCoV-19/Scotland/CVR2151/2020\|EPI_ISL_484564\|2020- | ···················· | ···················· | ···················· | ···················· | ···················· | ···················· | |
| E | 6203 | hCoV-19/Scotland/CVR2164/2020\|EPI_ISL_484565\|2020- | ···················· | ···················· | ···················· | ···················· | ···················· | ···················· | |
| E | 6204 | hCoV-19/Scotland/CVR2165/2020\|EPI_ISL_484566\|2020- | ···················· | ···················· | ···················· | ···················· | ···················· | ···················· | |
| E | 6205 | hCoV-19/Scotland/CVR2172/2020\|EPI_ISL_484567\|2020- | ···················· | ···················· | ···················· | ···················· | ···················· | ···················· | |
| E | 6206 | hCoV-19/Scotland/CVR2184/2020\|EPI_ISL_484568\|2020- | ···················· | ···················· | ···················· | ···················· | ···················· | ···················· | |
| E | 6207 | hCoV-19/Scotland/CVR2189/2020\|EPI_ISL_484569\|2020- | ···················· | ···················· | ···················· | ···················· | ···················· | ···················· | |
| E | 6208 | hCoV-19/Scotland/CVR2287/2020\|EPI_ISL_484570\|2020- | ···················· | ·A·················· | ···················· | ···················· | ···················· | ···················· | |
| E | 6209 | hCoV-19/Scotland/CVR2290/2020\|EPI_ISL_484571\|2020- | ···················· | ·A·················· | ···················· | ···················· | ···················· | ···················· | |
| E | 6210 | hCoV-19/Scotland/CVR2292/2020\|EPI_ISL_484572\|2020- | ···················· | ···················A | ···················· | ···················· | ···················· | ···················· | 1 |
| E | 6211 | hCoV-19/Scotland/CVR2296/2020\|EPI_ISL_484573\|2020- | ···················· | ···················· | ···················· | ···················· | ···················· | ···················· | |
| E | 6212 | hCoV-19/Germany/BW-UT-001/2020\|EPI_ISL_485809\|2020 | ···················· | ···················· | ···················· | ···················· | ···················· | ···················· | |
| E | 6213 | hCoV-19/Germany/BW-UT-002/2020\|EPI_ISL_485810\|2020 | ···················· | ···················· | ···················· | ···················· | ···················· | ···················· | |
| E | 6214 | hCoV-19/USA/VA-DCLS-0494/2020\|EPI_ISL_485814\|2020- | ···················· | ···················· | ···················· | ···················· | ···················· | ···················· | |
| E | 6215 | hCoV-19/USA/VA-DCLS-0498/2020\|EPI_ISL_485815\|2020- | ···················· | ···················· | ···················· | ···················· | ···················· | ···················· | |
| E | 6216 | hCoV-19/USA/VA-DCLS-0499/2020\|EPI_ISL_485816\|2020- | ···················· | ···················· | ···················· | ···················· | ···················· | ···················· | |

| | | | | | | | |
|---|---|---|---|---|---|---|---|
| E | 6217 hCoV-19/USA/VA-DCLS-0503/2020\|EPI_ISL_485817\|2020- | .................... | .................... | .................... | .................... | .................... | .................... |
| E | 6218 hCoV-19/USA/VA-DCLS-0506/2020\|EPI_ISL_485818\|2020- | .................... | .................... | .................... | .................... | .................... | .................... |
| E | 6219 hCoV-19/USA/VA-DCLS-0507/2020\|EPI_ISL_485819\|2020- | .................... | .................... | .................... | .................... | .................... | .................... |
| E | 6220 hCoV-19/USA/VA-DCLS-0509/2020\|EPI_ISL_485820\|2020- | .................... | .................... | .................... | .................... | .................... | .................... |
| E | 6221 hCoV-19/USA/VA-DCLS-0510/2020\|EPI_ISL_485821\|2020- | .................... | .................... | .................... | .................... | .................... | .................... |
| E | 6222 hCoV-19/USA/VA-DCLS-0511/2020\|EPI_ISL_485822\|2020- | .................... | .................... | .................... | .................... | .................... | .................... |
| E | 6223 hCoV-19/USA/VA-DCLS-0512/2020\|EPI_ISL_485823\|2020- | .................... | .................... | .................... | .................... | .................... | .................... |
| E | 6224 hCoV-19/USA/VA-DCLS-0513/2020\|EPI_ISL_485824\|2020- | .................... | .................... | .................... | .................... | .................... | .................... |
| E | 6225 hCoV-19/USA/VA-DCLS-0523/2020\|EPI_ISL_485825\|2020- | .................... | .................... | .................... | .................... | .................... | .................... |
| E | 6226 hCoV-19/USA/VA-DCLS-0524/2020\|EPI_ISL_485826\|2020- | .................... | .................... | .................... | .................... | .................... | .................... |
| E | 6227 hCoV-19/USA/VA-DCLS-0525/2020\|EPI_ISL_485827\|2020- | .................... | .................... | .................... | .................... | .................... | .................... |
| E | 6228 hCoV-19/USA/VA-DCLS-0526/2020\|EPI_ISL_485828\|2020- | .................... | .................... | .................... | .................... | .................... | .................... |
| E | 6229 hCoV-19/USA/VA-DCLS-0527/2020\|EPI_ISL_485829\|2020- | .................... | .................... | .................... | .................... | .................... | .................... |
| E | 6230 hCoV-19/USA/VA-DCLS-0528/2020\|EPI_ISL_485830\|2020- | .................... | .................... | .................... | .................... | .................... | .................... |
| E | 6231 hCoV-19/USA/VA-DCLS-0529/2020\|EPI_ISL_485831\|2020- | .................... | .................... | .................... | .................... | .................... | .................... |
| E | 6232 hCoV-19/USA/VA-DCLS-0530/2020\|EPI_ISL_485832\|2020- | .................... | .................... | .................... | .................... | .................... | .................... |
| E | 6233 hCoV-19/USA/VA-DCLS-0531/2020\|EPI_ISL_485833\|2020- | .................... | .................... | .................... | .................... | .................... | .................... |
| E | 6234 hCoV-19/USA/VA-DCLS-0533/2020\|EPI_ISL_485834\|2020- | .................... | .................... | .................... | .................... | .................... | .................... |
| E | 6235 hCoV-19/USA/VA-DCLS-0534/2020\|EPI_ISL_485835\|2020- | .................... | .................... | .................... | .................... | .................... | .................... |
| E | 6236 hCoV-19/USA/VA-DCLS-0535/2020\|EPI_ISL_485836\|2020- | .................... | .................... | .................... | .................... | .................... | .................... |
| E | 6237 hCoV-19/USA/VA-DCLS-0536/2020\|EPI_ISL_485837\|2020- | .................... | .................... | .................... | .................... | .................... | .................... |
| E | 6238 hCoV-19/USA/VA-DCLS-0537/2020\|EPI_ISL_485838\|2020- | .................... | .................... | .................... | .................... | .................... | .................... |
| E | 6239 hCoV-19/USA/VA-DCLS-0538/2020\|EPI_ISL_485839\|2020- | .................... | .................... | .................... | .................... | .................... | .................... |
| E | 6240 hCoV-19/USA/VA-DCLS-0539/2020\|EPI_ISL_485840\|2020- | .................... | .................... | .................... | .................... | .................... | .................... |
| E | 6241 hCoV-19/USA/VA-DCLS-0541/2020\|EPI_ISL_485842\|2020- | .................... | .................... | .................... | .................... | .................... | .................... |
| E | 6242 hCoV-19/USA/VA-DCLS-0542/2020\|EPI_ISL_485843\|2020- | .................... | .................... | .................... | .................... | .................... | .................... |
| E | 6243 hCoV-19/USA/VA-DCLS-0543/2020\|EPI_ISL_485844\|2020- | .................... | .................... | .................... | .................... | .................... | .................... |
| E | 6244 hCoV-19/USA/VA-DCLS-0544/2020\|EPI_ISL_485845\|2020- | .................... | .................... | .................... | .................... | .................... | .................... |
| E | 6245 hCoV-19/USA/VA-DCLS-0545/2020\|EPI_ISL_485846\|2020- | .................... | .................... | .................... | .................... | .................... | .................... |
| E | 6246 hCoV-19/USA/VA-DCLS-0546/2020\|EPI_ISL_485847\|2020- | .................... | .................... | .................... | .................... | .................... | .................... |
| E | 6247 hCoV-19/USA/VA-DCLS-0547/2020\|EPI_ISL_485848\|2020- | .................... | .................... | .................... | .................... | .................... | .................... |
| E | 6248 hCoV-19/USA/VA-DCLS-0548/2020\|EPI_ISL_485849\|2020- | .................... | .................... | .................... | .................... | .................... | .................... |
| E | 6249 hCoV-19/USA/VA-DCLS-0549/2020\|EPI_ISL_485850\|2020- | .................... | .................... | .................... | .................... | .................... | .................... |
| E | 6250 hCoV-19/USA/VA-DCLS-0550/2020\|EPI_ISL_485851\|2020- | .................... | .................... | .................... | .................... | .................... | .................... |
| E | 6251 hCoV-19/USA/VA-DCLS-0551/2020\|EPI_ISL_485852\|2020- | .................... | .................... | .................... | .................... | .................... | .................... |
| E | 6252 hCoV-19/USA/VA-DCLS-0552/2020\|EPI_ISL_485853\|2020- | .................... | .................... | .................... | .................... | .................... | .................... |
| E | 6253 hCoV-19/USA/VA-DCLS-0553/2020\|EPI_ISL_485854\|2020- | .................... | .................... | .................... | .................... | .................... | .................... |
| E | 6254 hCoV-19/USA/VA-DCLS-0554/2020\|EPI_ISL_485855\|2020- | .................... | .................... | .................... | .................... | .................... | .................... |
| E | 6255 hCoV-19/USA/VA-DCLS-0555/2020\|EPI_ISL_485856\|2020- | .................... | .................... | .................... | .................... | .................... | .................... |
| E | 6256 hCoV-19/USA/VA-DCLS-0556/2020\|EPI_ISL_485857\|2020- | .................... | .................... | .................... | .................... | .................... | .................... |
| E | 6257 hCoV-19/USA/VA-DCLS-0557/2020\|EPI_ISL_485858\|2020- | .................... | .................... | .................... | .................... | .................... | .................... |
| E | 6258 hCoV-19/USA/VA-DCLS-0558/2020\|EPI_ISL_485859\|2020- | .................... | .................... | .................... | .................... | .................... | .................... |
| E | 6259 hCoV-19/USA/VA-DCLS-0559/2020\|EPI_ISL_485860\|2020- | .................... | .................... | .................... | .................... | .................... | .................... |
| E | 6260 hCoV-19/USA/LA-GBCL-26544531/2020\|EPI_ISL_485885\|2 | .................... | .................... | .................... | .................... | .................... | .................... |
| E | 6261 hCoV-19/USA/LA-GBCL-26544562/2020\|EPI_ISL_485888\|2 | .................... | .................... | .................... | .................... | .................... | .................... |
| E | 6262 hCoV-19/USA/LA-GBCL-26544578/2020\|EPI_ISL_485889\|2 | .................... | .................... | .................... | .................... | .................... | .................... |
| E | 6263 hCoV-19/USA/LA-GBCL-26556571/2020\|EPI_ISL_485890\|2 | .................... | .................... | .................... | .................... | .................... | .................... |
| E | 6264 hCoV-19/USA/LA-GBCL-26629191/2020\|EPI_ISL_485892\|2 | .................... | .................... | .................... | .................... | .................... | .................... |
| E | 6265 hCoV-19/USA/LA-GBCL-26629251/2020\|EPI_ISL_485895\|2 | .................... | .................... | .................... | .................... | .................... | .................... |
| E | 6266 hCoV-19/USA/LA-GBCL-26629256/2020\|EPI_ISL_485896\|2 | .................... | .................... | .................... | .................... | .................... | .................... |
| E | 6267 hCoV-19/USA/LA-GBCL-26674724/2020\|EPI_ISL_485897\|2 | .................... | .................... | .................... | .................... | .................... | .................... |
| E | 6268 hCoV-19/England/NORT-29741F/2020\|EPI_ISL_488018\|20 | .................... | .................... | .................... | .................... | .................... | .................... |
| E | 6269 hCoV-19/England/NORT-2982D8/2020\|EPI_ISL_488020\|20 | .................... | .................... | .................... | .................... | .................... | .................... |
| E | 6270 hCoV-19/England/NORT-2976DD/2020\|EPI_ISL_488021\|20 | .................... | .................... | .................... | .................... | .................... | .................... |
| E | 6271 hCoV-19/England/NORT-29789B/2020\|EPI_ISL_488022\|20 | .................... | .................... | .................... | .................... | .................... | .................... |
| E | 6272 hCoV-19/England/NORT-2978D7/2020\|EPI_ISL_488023\|20 | .................... | .................... | .................... | .................... | .................... | .................... |
| E | 6273 hCoV-19/England/NORT-296B75/2020\|EPI_ISL_488025\|20 | .................... | .................... | .................... | .................... | .................... | .................... |
| E | 6274 hCoV-19/England/NORT-2985DF/2020\|EPI_ISL_488026\|20 | .................... | .................... | .................... | .................... | .................... | .................... |
| E | 6275 hCoV-19/England/NORT-29743D/2020\|EPI_ISL_488028\|20 | .................... | .................... | .................... | .................... | .................... | .................... |
| E | 6276 hCoV-19/England/NORT-297707/2020\|EPI_ISL_488030\|20 | .................... | .................... | .................... | .................... | .................... | .................... |
| E | 6277 hCoV-19/England/NORT-297664/2020\|EPI_ISL_488036\|20 | .................... | .................... | .................... | .................... | .................... | .................... |
| E | 6278 hCoV-19/England/NORT-2964D4/2020\|EPI_ISL_488038\|20 | .................... | .................... | .................... | .................... | .................... | .................... |
| E | 6279 hCoV-19/England/NORT-297C62/2020\|EPI_ISL_488060\|20 | .................... | .................... | .................... | .................... | .................... | .................... |
| E | 6280 hCoV-19/England/NORT-29D965/2020\|EPI_ISL_488061\|20 | .................... | .................... | .................... | .................... | .................... | .................... |
| E | 6281 hCoV-19/England/NORT-29C7F3/2020\|EPI_ISL_488063\|20 | .................... | .................... | .................... | .................... | .................... | .................... |
| E | 6282 hCoV-19/England/NORT-29E59D/2020\|EPI_ISL_488064\|20 | .................... | .................... | .................... | .................... | .................... | .................... |
| E | 6283 hCoV-19/England/NORT-29E2D2/2020\|EPI_ISL_488065\|20 | .................... | .................... | .................... | .................... | .................... | .................... |
| E | 6284 hCoV-19/England/NORT-29D200/2020\|EPI_ISL_488066\|20 | .................... | .................... | .................... | .................... | .................... | .................... |
| E | 6285 hCoV-19/England/NORT-29EAF8/2020\|EPI_ISL_488067\|20 | .................... | .................... | .................... | .................... | .................... | .................... |
| E | 6286 hCoV-19/England/NORT-297D14/2020\|EPI_ISL_488068\|20 | .................... | .................... | .................... | .................... | .................... | .................... |
| E | 6287 hCoV-19/England/NORT-29CE67/2020\|EPI_ISL_488069\|20 | .................... | .................... | .................... | .................... | .................... | .................... |
| E | 6288 hCoV-19/England/NORT-298706/2020\|EPI_ISL_488070\|20 | .................... | .................... | .................... | .................... | .................... | .................... |
| E | 6289 hCoV-19/England/NORT-29E15D/2020\|EPI_ISL_488071\|20 | .................... | .................... | .................... | .................... | .................... | .................... |
| E | 6290 hCoV-19/England/NORT-29800B/2020\|EPI_ISL_488075\|20 | .................... | .................... | .................... | .................... | .................... | .................... |
| E | 6291 hCoV-19/England/NORT-29D9DE/2020\|EPI_ISL_488076\|20 | .................... | .................... | .................... | .................... | .................... | .................... |
| E | 6292 hCoV-19/England/NORT-298250/2020\|EPI_ISL_488077\|20 | .................... | .................... | .................... | .................... | .................... | .................... |
| E | 6293 hCoV-19/England/NORT-2985B1/2020\|EPI_ISL_488078\|20 | .................... | .................... | .................... | .................... | .................... | .................... |
| E | 6294 hCoV-19/England/NORT-29DEFD/2020\|EPI_ISL_488080\|20 | .................... | .................... | .................... | .................... | .................... | .................... |
| E | 6295 hCoV-19/England/NORT-29DC20/2020\|EPI_ISL_488081\|20 | .................... | .................... | .................... | .................... | .................... | .................... |
| E | 6296 hCoV-19/England/NORT-29DC7B/2020\|EPI_ISL_488084\|20 | .................... | .................... | .................... | .................... | .................... | .................... |

| | | | | | | | | |
|---|---|---|---|---|---|---|---|---|
| E | 6297 | hCoV-19/England/NORT-297D23/2020\|EPI_ISL_488085\|20 | ···················· | ···················· | ···················· | ···················· | ···················· | ···················· |
| E | 6298 | hCoV-19/England/NORT-296DE8/2020\|EPI_ISL_488087\|20 | ···················· | ···················· | ···················· | ···················· | ···················· | ···················· |
| E | 6299 | hCoV-19/England/NORT-29D895/2020\|EPI_ISL_488088\|20 | ···················· | ···················· | ···················· | ···················· | ···················· | ···················· |
| E | 6300 | hCoV-19/England/NORT-297F1E/2020\|EPI_ISL_488089\|20 | ···················· | ···················· | ···················· | ···················· | ···················· | ···················· |
| E | 6301 | hCoV-19/England/NORT-297D6F/2020\|EPI_ISL_488090\|20 | ···················· | ···················· | ···················· | ···················· | ···················· | ···················· |
| E | 6302 | hCoV-19/England/NORT-29E0E7/2020\|EPI_ISL_488091\|20 | ···················· | ···················· | ···················· | ···················· | ···················· | ···················· |
| E | 6303 | hCoV-19/England/NORT-29EE1A/2020\|EPI_ISL_488092\|20 | ···················· | ···················· | ···················· | ···················· | ···················· | ···················· |
| E | 6304 | hCoV-19/England/NORT-297D41/2020\|EPI_ISL_488093\|20 | ···················· | ···················· | ···················· | ···················· | ···················· | ···················· |
| E | 6305 | hCoV-19/England/NORT-29EA43/2020\|EPI_ISL_488095\|20 | ···················· | ···················· | ···················· | ···················· | ···················· | ···················· |
| E | 6306 | hCoV-19/England/NORT-29827E/2020\|EPI_ISL_488096\|20 | ···················· | ···················· | ···················· | ···················· | ···················· | ···················· |
| E | 6307 | hCoV-19/England/NORT-29C76C/2020\|EPI_ISL_488099\|20 | ···················· | ···················· | ···················· | ···················· | ···················· | ···················· |
| E | 6308 | hCoV-19/Northern Ireland/NIRE-103682/2020\|EPI_ISL_ | ···················· | ···················· | ···················· | ···················· | ···················· | ···················· |
| E | 6309 | hCoV-19/Northern Ireland/NIRE-10444B/2020\|EPI_ISL_ | ···················· | ···················· | ···················· | ···················· | ···················· | ···················· |
| E | 6310 | hCoV-19/Northern Ireland/NIRE-FD130/2020\|EPI_ISL_4 | ···················· | ···················· | ···················· | ···················· | ···················· | ···················· |
| E | 6311 | hCoV-19/Northern Ireland/NIRE-FD1A9/2020\|EPI_ISL_4 | ···················· | ···················· | ···················· | ···················· | ···················· | ···················· |
| E | 6312 | hCoV-19/Northern Ireland/NIRE-FD640/2020\|EPI_ISL_4 | ···················· | ···················· | ···················· | ···················· | ···················· | ···················· |
| E | 6313 | hCoV-19/Northern Ireland/NIRE-103664/2020\|EPI_ISL_ | ···················· | ···················· | ···················· | ···················· | ···················· | ···················· |
| E | 6314 | hCoV-19/Northern Ireland/NIRE-100184/2020\|EPI_ISL_ | ···················· | ···················· | ···················· | ···················· | ···················· | ···················· |
| E | 6315 | hCoV-19/Northern Ireland/NIRE-103154/2020\|EPI_ISL_ | ···················· | ···················· | ···················· | ···················· | ···················· | ···················· |
| E | 6316 | hCoV-19/Northern Ireland/NIRE-FCE2B/2020\|EPI_ISL_4 | ···················· | ···················· | ···················· | ···················· | ···················· | ···················· |
| E | 6317 | hCoV-19/Northern Ireland/NIRE-103400/2020\|EPI_ISL_ | ···················· | ···················· | ···················· | ···················· | ···················· | ···················· |
| E | 6318 | hCoV-19/Northern Ireland/NIRE-FD18B/2020\|EPI_ISL_4 | ···················· | ···················· | ···················· | ···················· | ···················· | ···················· |
| E | 6319 | hCoV-19/Northern Ireland/NIRE-1032BB/2020\|EPI_ISL_ | ···················· | ···················· | ···················· | ···················· | ···················· | ···················· |
| E | 6320 | hCoV-19/Northern Ireland/NIRE-1031AF/2020\|EPI_ISL_ | ···················· | ···················· | ···················· | ···················· | ···················· | ···················· |
| E | 6321 | hCoV-19/Northern Ireland/NIRE-1035B2/2020\|EPI_ISL_ | ···················· | ···················· | ···················· | ···················· | ···················· | ···················· |
| E | 6322 | hCoV-19/Northern Ireland/NIRE-FD0E8/2020\|EPI_ISL_4 | ···················· | ···················· | ···················· | ···················· | ···················· | ···················· |
| E | 6323 | hCoV-19/Northern Ireland/NIRE-103691/2020\|EPI_ISL_ | ···················· | ···················· | ···················· | ···················· | ···················· | ···················· |
| E | 6324 | hCoV-19/Northern Ireland/NIRE-FCE94/2020\|EPI_ISL_4 | ···················· | ···················· | ···················· | ···················· | ···················· | ···················· |
| E | 6325 | hCoV-19/England/CAMB-1ACEED/2020\|EPI_ISL_489390\|20 | ···················· | ···················· | ···················· | ···················· | ···················· | ···················· |
| E | 6326 | hCoV-19/Scotland/GCVR-171ADE/2020\|EPI_ISL_489391\|2 | ···················· | ·A·················· | ···················· | ···················· | ···················· | ···················· |
| E | 6327 | hCoV-19/Scotland/GCVR-171AB0/2020\|EPI_ISL_489392\|2 | ···················· | ···················· | ··A················· | ···················· | ···················· | ···················· |
| E | 6328 | hCoV-19/England/CAMB-1ACD95/2020\|EPI_ISL_489394\|20 | ···················· | ···················· | ···················· | ···················· | ···················· | ···················· |
| E | 6329 | hCoV-19/England/CAMB-1ACE56/2020\|EPI_ISL_489395\|20 | ···················· | ···················· | ···················· | ···················· | ···················· | ···················· |
| E | 6330 | hCoV-19/Scotland/GCVR-1719D1/2020\|EPI_ISL_489396\|2 | ···················· | ···················· | ···················· | ···················· | ···················· | ···················· |
| E | 6331 | hCoV-19/England/CAMB-1AD57E/2020\|EPI_ISL_489397\|20 | ···················· | ···················· | ···················· | ···················· | ···················· | ···················· |
| E | 6332 | hCoV-19/England/CVR3937/2020\|EPI_ISL_490330\|2020-0 | ···················· | ···················· | ···················· | ···················· | ···················· | ···················· |
| E | 6334 | hCoV-19/England/LIVE-A1793/2020\|EPI_ISL_490337\|202 | ···················· | ···················· | ···················· | ···················· | ···················· | ···················· |
| E | 6335 | hCoV-19/England/LIVE-A17B1/2020\|EPI_ISL_490338\|202 | ···················· | ···················· | ···················· | ···················· | ···················· | ···················· |
| E | 6336 | hCoV-19/England/LIVE-A17EE/2020\|EPI_ISL_490339\|202 | ···················· | ···················· | ···················· | ···················· | ···················· | ···················· |
| E | 6337 | hCoV-19/England/LIVE-A18EB/2020\|EPI_ISL_490343\|202 | ···················· | ···················· | ···················· | ···················· | ···················· | ···················· |
| E | 6338 | hCoV-19/England/LIVE-A1915/2020\|EPI_ISL_490345\|202 | ···················· | ···················· | ···················· | ···················· | ···················· | ···················· |
| E | 6339 | hCoV-19/England/LIVE-A1924/2020\|EPI_ISL_490346\|202 | ···················· | ···················· | ···················· | ···················· | ···················· | ···················· |
| E | 6340 | hCoV-19/England/LIVE-A1933/2020\|EPI_ISL_490347\|202 | ···················· | ···················· | ···················· | ···················· | ···················· | ···················· |
| E | 6341 | hCoV-19/England/LIVE-A1942/2020\|EPI_ISL_490348\|202 | ···················· | ···················· | ···················· | ···················· | ···················· | ···················· |
| E | 6342 | hCoV-19/England/LIVE-A1FAA/2020\|EPI_ISL_490371\|202 | ···················· | ···················· | ···················· | ···················· | ···················· | ···················· |
| E | 6345 | hCoV-19/England/LIVE-A212B/2020\|EPI_ISL_490377\|202 | ···················· | ···················· | ···················· | ···················· | ···················· | ···················· |
| E | 6346 | hCoV-19/England/LIVE-A2167/2020\|EPI_ISL_490378\|202 | ···················· | ···················· | ···················· | ···················· | ···················· | ···················· |
| E | 6348 | hCoV-19/England/LIVE-A2404/2020\|EPI_ISL_490380\|202 | ···················· | ···················· | ···················· | ···················· | ···················· | ···················· |
| E | 6349 | hCoV-19/England/LIVE-A2431/2020\|EPI_ISL_490382\|202 | ···················· | ···················· | ···················· | ···················· | ···················· | ···················· |
| E | 6350 | hCoV-19/England/LIVE-A3F4E/2020\|EPI_ISL_490398\|202 | ···················· | ···················· | ···················· | ···················· | ···················· | ···················· |
| E | 6351 | hCoV-19/Scotland/EDB2274/2020\|EPI_ISL_491660\|2020- | ···················· | ···················· | ···················· | ···················· | ···················· | ···················· |
| E | 6352 | hCoV-19/Scotland/EDB1840/2020\|EPI_ISL_491661\|2020- | ···················· | ···················· | ···················· | ···················· | ···················· | ···················· |
| E | 6353 | hCoV-19/Scotland/EDB1242/2020\|EPI_ISL_491662\|2020- | ···················· | ···················· | ···················· | ···················· | ···················· | ···················· |
| E | 6354 | hCoV-19/Scotland/EDB2124/2020\|EPI_ISL_491663\|2020- | ···················· | ···················· | ···················· | ···················· | ···················· | ···················· |
| E | 6355 | hCoV-19/Scotland/EDB1942/2020\|EPI_ISL_491664\|2020- | ···················· | ·A·················· | ···················· | ···················· | ···················· | ···················· |
| E | 6356 | hCoV-19/Scotland/EDB2457/2020\|EPI_ISL_491665\|2020- | ···················· | ·A·················· | ···················· | ···················· | ···················· | ···················· |
| E | 6357 | hCoV-19/Scotland/EDB1408/2020\|EPI_ISL_491667\|2020- | ···················· | ·A·················· | ···················· | ···················· | ···················· | ···················· |
| E | 6358 | hCoV-19/Scotland/EDB698/2020\|EPI_ISL_491668\|2020-0 | ···················· | ···················· | ···················· | ···················· | ···················· | ···················· |
| E | 6359 | hCoV-19/Scotland/EDB1916/2020\|EPI_ISL_491669\|2020- | ···················· | ·A·················· | ···················· | ···················· | ···················· | ···················· |
| E | 6360 | hCoV-19/Scotland/EDB1837/2020\|EPI_ISL_491670\|2020- | ···················· | ···················· | ···················· | ···················· | ···················· | ···················· |
| E | 6361 | hCoV-19/Scotland/EDB1473/2020\|EPI_ISL_491672\|2020- | ···················· | ···················· | ···················· | ···················· | ···················· | ···················· |
| E | 6362 | hCoV-19/Scotland/EDB2481/2020\|EPI_ISL_491674\|2020- | ···················· | ···················· | ···················· | ···················· | ···················· | ···················· |
| E | 6363 | hCoV-19/Scotland/EDB1272/2020\|EPI_ISL_491675\|2020- | ···················· | ···················· | ···················· | ···················· | ···················· | ···················· |
| E | 6364 | hCoV-19/Scotland/EDB3776/2020\|EPI_ISL_491676\|2020- | ···················· | ···················· | ···················· | ···················· | ···················· | ···················· |
| E | 6365 | hCoV-19/Scotland/EDB2145/2020\|EPI_ISL_491677\|2020- | ···················· | ·A·················· | ···················· | ···················· | ···················· | ···················· |
| E | 6366 | hCoV-19/Scotland/EDB2546/2020\|EPI_ISL_491678\|2020- | ···················· | ···················· | ···················· | ···················· | ···················· | ···················· |
| E | 6367 | hCoV-19/Scotland/EDB6448/2020\|EPI_ISL_491679\|2020- | ···················· | ···················· | ···················· | ···················· | ···················· | ···················· |
| E | 6368 | hCoV-19/Scotland/EDB1887/2020\|EPI_ISL_491691\|2020- | ···················· | ···················· | ···················· | ···················· | ···················· | ···················· |
| E | 6369 | hCoV-19/Latvia/055/2020\|EPI_ISL_492990\|2020-07-08 | ···················· | ···················· | ···················· | ···················· | ···················· | ···················· |
| E | 6370 | hCoV-19/Latvia/056/2020\|EPI_ISL_492991\|2020-07-08 | ···················· | ···················· | ···················· | ···················· | ···················· | ···················· |
| E | 6371 | hCoV-19/Latvia/057/2020\|EPI_ISL_492992\|2020-07-07 | ···················· | ···················· | ···················· | ···················· | ···················· | ···················· |
| E | 6372 | hCoV-19/Latvia/058/2020\|EPI_ISL_492993\|2020-07-08 | ···················· | ···················· | ···················· | ···················· | ···················· | ···················· |
| E | 6373 | hCoV-19/Latvia/059/2020\|EPI_ISL_492994\|2020-07-08 | ···················· | ···················· | ···················· | ···················· | ···················· | ···················· |
| E | 6374 | hCoV-19/Latvia/060/2020\|EPI_ISL_492995\|2020-07-07 | ···················· | ···················· | ···················· | ···················· | ···················· | ···················· |
| E | 6375 | hCoV-19/Latvia/061/2020\|EPI_ISL_492996\|2020-07-07 | ···················· | ···················· | ···················· | ···················· | ···················· | ···················· |
| E | 6376 | hCoV-19/Latvia/062/2020\|EPI_ISL_492997\|2020-07-07 | ···················· | ···················· | ···················· | ···················· | ···················· | ···················· |
| E | 6377 | hCoV-19/Latvia/063/2020\|EPI_ISL_492998\|2020-07-03 | ···················· | ···················· | ···················· | ···················· | ···················· | ···················· |
| E | 6378 | hCoV-19/Latvia/064/2020\|EPI_ISL_492999\|2020-07-09 | ···················· | ···················· | ···················· | ···················· | ···················· | ···················· |
| E | 6379 | hCoV-19/India/TG-CCMB-M359/2020\|EPI_ISL_495200\|202 | ···················· | ···················· | ···················· | ···················· | ···················· | ···················· |
| E | 6380 | hCoV-19/India/TG-CCMB-M361/2020\|EPI_ISL_495201\|202 | ···················· | ···················· | ···················· | ···················· | ···················· | ···················· |

| | | | | | | | | |
|---|---|---|---|---|---|---|---|---|
| E | 6381 hCoV-19/India/TG-CCMB-M363/2020\|EPI_ISL_495202\|202 | ··················T· | ···················· | ···················· | ···················· | ···················· | ···················· | 1 |
| E | 6382 hCoV-19/India/TG-CCMB-M437/2020\|EPI_ISL_495203\|202 | ···················· | ···················· | ···················· | ···················· | ···················· | ···················· | |
| E | 6383 hCoV-19/India/TG-CCMB-M441/2020\|EPI_ISL_495204\|202 | ···················· | ···················· | ···················· | ···················· | ···················· | ···················· | |
| E | 6386 hCoV-19/India/TG-CCMB-M456/2020\|EPI_ISL_495207\|202 | ···················· | ···················· | ···················· | ···················· | ···················· | ···················· | |
| E | 6387 hCoV-19/India/TG-CCMB-M458/2020\|EPI_ISL_495208\|202 | ···················· | ···················· | ···················· | ···················· | ···················· | ···················· | |
| E | 6388 hCoV-19/India/TG-CCMB-M465/2020\|EPI_ISL_495209\|202 | ···················· | ···················· | ···················· | ···················· | ···················· | ···················· | |
| E | 6389 hCoV-19/India/TG-CCMB-M466/2020\|EPI_ISL_495210\|202 | ···················· | ···················· | ···················· | ···················· | ···················· | ···················· | |
| E | 6390 hCoV-19/India/TG-CCMB-M471/2020\|EPI_ISL_495211\|202 | ···················· | ···················· | ···················· | ···················· | ···················· | ···················· | |
| E | 6391 hCoV-19/India/TG-CCMB-M489/2020\|EPI_ISL_495212\|202 | ···················· | ···················· | ···················· | ···················· | ···················· | ···················· | |
| E | 6392 hCoV-19/India/TG-CCMB-M499/2020\|EPI_ISL_495213\|202 | ···················· | ···················· | ···················· | ···················· | ···················· | ···················· | |
| E | 6393 hCoV-19/India/TG-CCMB-M5/2020\|EPI_ISL_495214\|2020- | ···················· | ···················· | ···················· | ···················· | ···················N | ···················· | 1 |
| E | 6396 hCoV-19/India/TG-CCMB-M588/2020\|EPI_ISL_495217\|202 | ···················· | ···················· | ···················· | ···················· | ···················· | ···················· | |
| E | 6397 hCoV-19/India/TG-CCMB-M64/2020\|EPI_ISL_495218\|2020 | ···················· | ···················· | ···················· | ···················· | ···················· | ···················· | |
| E | 6399 hCoV-19/India/TG-CCMB-M652/2020\|EPI_ISL_495220\|202 | ···················· | ···················· | ···················· | ···················· | ···················· | ···················· | |
| E | 6400 hCoV-19/India/TG-CCMB-M66/2020\|EPI_ISL_495221\|2020 | ···················· | ···················· | ···················· | ···················· | ···················· | ···················· | |
| E | 6401 hCoV-19/England/HECH-2599D64/2020\|EPI_ISL_482078\|2 | ···················· | ···················· | ···················· | ···················· | ···················· | ···················· | |
| E | 6402 hCoV-19/England/HECH-2599C76/2020\|EPI_ISL_482079\|2 | ···················· | ···················· | ···················· | ···················· | ···················· | ···················· | |
| E | 6403 hCoV-19/England/HECH-2599CEF/2020\|EPI_ISL_482081\|2 | ···················· | ·············A······ | ···················· | ···················· | ···················· | ···················· | |
| E | 6404 hCoV-19/Northern Ireland/NIRE-104180/2020\|EPI_ISL_ | ···················· | ···················· | ···················· | ···················· | ···················· | ···················· | |
| E | 6405 hCoV-19/England/HECH-2599E52/2020\|EPI_ISL_482084\|2 | ···················· | ···················· | ···················· | ···················· | ···················· | ···················· | |
| E | 6406 hCoV-19/England/HECH-2599CC1/2020\|EPI_ISL_482085\|2 | ···················· | ···················· | ···················· | ···················· | ···················· | ···················· | |
| E | 6407 hCoV-19/England/HECH-259A000/2020\|EPI_ISL_482086\|2 | ···················· | ···················· | ···················· | ···················· | ···················· | ···················· | |
| E | 6408 hCoV-19/England/HECH-2599CB2/2020\|EPI_ISL_482088\|2 | ···················· | ···················· | ···················· | ···················· | ···················· | ···················· | |
| E | 6409 hCoV-19/England/HECH-2599E43/2020\|EPI_ISL_482089\|2 | ···················· | ···················· | ···················· | ···················· | ···················· | ···················· | |
| E | 6410 hCoV-19/England/HECH-2599EDA/2020\|EPI_ISL_482090\|2 | ···················· | ···················· | ···················· | ···················· | ···················· | ···················· | |
| E | 6411 hCoV-19/England/HECH-2599C94/2020\|EPI_ISL_482092\|2 | ···················· | ···················· | ···················· | ···················· | ···················· | ···················· | |
| E | 6412 hCoV-19/England/HECH-259A097/2020\|EPI_ISL_482093\|2 | ···················· | ···················· | ···················· | ···················· | ···················· | ···················· | |
| E | 6413 hCoV-19/England/HECH-2599C85/2020\|EPI_ISL_482094\|2 | ···················· | ···················· | ···················· | ···················· | ···················· | ···················· | |
| E | 6414 hCoV-19/England/HECH-2599C67/2020\|EPI_ISL_482095\|2 | ···················· | ···················· | ···················· | ···················· | ···················· | ···················· | |
| E | 6415 hCoV-19/Northern Ireland/NIRE-1013D8/2020\|EPI_ISL_ | ···················· | ···················· | ···················· | ···················· | ···················· | ···················· | |
| E | 6416 hCoV-19/England/HECH-259A02E/2020\|EPI_ISL_482099\|2 | ···················· | ···················· | ···················· | ···················· | ···················· | ···················· | |
| E | 6417 hCoV-19/England/NORT-296313/2020\|EPI_ISL_488039\|20 | ···················· | ···················· | ···················· | ···················· | ···················· | ···················· | |
| E | 6418 hCoV-19/England/NORT-296234/2020\|EPI_ISL_488040\|20 | ···················· | ···················A | ···················· | ···················· | ···················· | ···················· | 1 |
| E | 6419 hCoV-19/England/NORT-29639B/2020\|EPI_ISL_488042\|20 | ···················· | ···················· | ···················· | ···················· | ···················· | ···················· | |
| E | 6420 hCoV-19/England/NORT-296243/2020\|EPI_ISL_488044\|20 | ···················· | ···················A | ···················· | ···················· | ···················· | ···················· | 1 |
| E | 6421 hCoV-19/England/NORT-2966B0/2020\|EPI_ISL_488046\|20 | ···················· | ···················· | ···················· | ···················· | ···················· | ···················· | |
| E | 6422 hCoV-19/England/NORT-2963AA/2020\|EPI_ISL_488047\|20 | ···················· | ···················· | ···················· | ···················· | ···················· | ···················· | |
| E | 6423 hCoV-19/England/NORT-296771/2020\|EPI_ISL_488048\|20 | ···················· | ···················· | ···················· | ···················· | ···················· | ···················· | |
| E | 6424 hCoV-19/England/NORT-296322/2020\|EPI_ISL_488049\|20 | ···················· | ···················· | ···················· | ···················· | ···················· | ···················· | |
| E | 6425 hCoV-19/England/NORT-29F71E/2020\|EPI_ISL_488051\|20 | ···················· | ·A·················· | ···················· | ···················· | ···················· | ···················· | |
| E | 6426 hCoV-19/England/NORT-29D473/2020\|EPI_ISL_488053\|20 | ···················· | ···················· | ···················· | ···················· | ···················· | ···················· | |
| E | 6427 hCoV-19/England/NORT-295F98/2020\|EPI_ISL_488055\|20 | ···················· | ···················· | ···················· | ···················· | ···················· | ···················· | |
| E | 6428 hCoV-19/England/NORT-29C85A/2020\|EPI_ISL_488056\|20 | ···················· | ···················· | ···················· | ···················· | ···················· | ···················· | |
| E | 6429 hCoV-19/England/NORT-29ECD4/2020\|EPI_ISL_488057\|20 | ···················· | ···················A | ···················· | ···················· | ···················· | ···················· | 1 |
| E | 6430 hCoV-19/England/NORT-29E1C6/2020\|EPI_ISL_488058\|20 | ···················· | ···················· | ···················· | ···················· | ···················· | ···················· | |
| E | 6431 hCoV-19/England/BRIS-12F3E3/2020\|EPI_ISL_488200\|20 | ···················· | ···················· | ···················· | ···················· | ···················· | ···················· | |
| E | 6432 hCoV-19/England/BRIS-12F811/2020\|EPI_ISL_488201\|20 | ···················· | ···················· | ···················· | ···················· | ···················· | ···················· | |
| E | 6433 hCoV-19/England/BRIS-12F5ED/2020\|EPI_ISL_488202\|20 | ···················· | ···················· | ···················· | ···················· | ···················· | ···················· | |
| E | 6434 hCoV-19/England/BRIS-12EED4/2020\|EPI_ISL_488203\|20 | ···················· | ···················· | ···················· | ···················· | ···················· | ···················· | |
| E | 6435 hCoV-19/England/BRIS-12E681/2020\|EPI_ISL_488205\|20 | ···················· | ···················· | ···················· | ···················· | ···················· | ···················· | |
| E | 6436 hCoV-19/England/BRIS-12EBFB/2020\|EPI_ISL_488206\|20 | ···················· | ···················· | ···················· | ···················· | ···················· | ···················· | |
| E | 6437 hCoV-19/England/BRIS-12EC52/2020\|EPI_ISL_488207\|20 | ···················· | ···················· | ···················· | ···················· | ···················· | ···················· | |
| E | 6438 hCoV-19/England/BRIS-12EA0D/2020\|EPI_ISL_488208\|20 | ···················· | ···················· | ···················· | ···················· | ···················· | ···················· | |
| E | 6439 hCoV-19/England/BRIS-12E8A9/2020\|EPI_ISL_488209\|20 | ···················· | ·R·················· | ···················· | ···················· | ···················· | ···················· | |
| E | 6440 hCoV-19/England/BRIS-12E8F4/2020\|EPI_ISL_488210\|20 | ···················· | ···················· | ···················· | ···················· | ···················· | ···················· | |
| E | 6441 hCoV-19/England/BRIS-12E557/2020\|EPI_ISL_488213\|20 | ···················· | ···················· | ···················· | ···················· | ···················· | ···················· | |
| E | 6442 hCoV-19/England/BRIS-12F87B/2020\|EPI_ISL_488216\|20 | ···················· | ···················· | ···················· | ···················· | ···················· | ···················· | |
| E | 6443 hCoV-19/England/BRIS-12E8E5/2020\|EPI_ISL_488217\|20 | ···················· | ···················· | ···················· | ···················· | ···················· | ···················· | |
| E | 6444 hCoV-19/England/BRIS-12F152/2020\|EPI_ISL_488218\|20 | ···················· | ···················· | ···················· | ···················· | ···················· | ···················· | |
| E | 6445 hCoV-19/England/BRIS-12F5A1/2020\|EPI_ISL_488219\|20 | ···················· | ···················· | ···················· | ···················· | ···················· | ···················· | |
| E | 6446 hCoV-19/England/BRIS-12EF1D/2020\|EPI_ISL_488220\|20 | ···················· | ···················· | ···················· | ···················· | ···················· | ···················· | |
| E | 6447 hCoV-19/England/BRIS-12F635/2020\|EPI_ISL_488221\|20 | ···················· | ···················· | ···················· | ···················· | ···················· | ···················· | |
| E | 6448 hCoV-19/England/BRIS-12E5B1/2020\|EPI_ISL_488222\|20 | ···················· | ···················· | ···················· | ···················· | ···················· | ···················· | |
| E | 6449 hCoV-19/England/BRIS-12F705/2020\|EPI_ISL_488223\|20 | ···················· | ···················· | ···················· | ···················· | ···················· | ···················· | |
| E | 6450 hCoV-19/England/BRIS-12E4E1/2020\|EPI_ISL_488224\|20 | ···················· | ···················· | ···················· | ···················· | ···················· | ···················· | |
| E | 6451 hCoV-19/England/BRIS-12E78E/2020\|EPI_ISL_488225\|20 | ···················· | ···················· | ···················· | ···················· | ···················· | ···················· | |
| E | 6452 hCoV-19/England/BRIS-12F1DA/2020\|EPI_ISL_488226\|20 | ···················· | ···················· | ···················· | ···················· | ···················· | ···················· | |
| E | 6453 hCoV-19/England/BRIS-12E7BB/2020\|EPI_ISL_488227\|20 | ···················· | ···················· | ···················· | ···················· | ···················· | ···················· | |
| E | 6454 hCoV-19/England/BRIS-12F6DB/2020\|EPI_ISL_488228\|20 | ···················· | ···················· | ···················· | ···················· | ···················· | ···················· | |
| E | 6455 hCoV-19/England/BRIS-12F240/2020\|EPI_ISL_488230\|20 | ···················· | ···················· | ···················· | ···················· | ···················· | ···················· | |
| E | 6456 hCoV-19/England/BRIS-12EBDD/2020\|EPI_ISL_488231\|20 | ···················· | ···················· | ···················· | ···················· | ···················· | ···················· | |
| E | 6457 hCoV-19/England/BRIS-12F50B/2020\|EPI_ISL_488232\|20 | ···················· | ···················· | ···················· | ···················· | ···················· | ···················· | |
| E | 6458 hCoV-19/England/BRIS-12E575/2020\|EPI_ISL_488234\|20 | ···················· | ···················· | ···················· | ···················· | ···················· | ···················· | |
| E | 6459 hCoV-19/England/BRIS-12EA85/2020\|EPI_ISL_488236\|20 | ···················· | ···················· | ···················· | ···················· | ···················· | ···················· | |
| E | 6460 hCoV-19/England/BRIS-12F2B9/2020\|EPI_ISL_488237\|20 | ···················· | ···················A | ···················· | ···················· | ···················· | ···················· | 1 |
| E | 6461 hCoV-19/England/BRIS-12F2E6/2020\|EPI_ISL_488238\|20 | ···················· | ···················· | ···················· | ···················· | ···················· | ···················· | |
| E | 6462 hCoV-19/England/BRIS-12F116/2020\|EPI_ISL_488239\|20 | ···················· | ···················· | ···················· | ···················· | ···················· | ···················· | |
| E | 6463 hCoV-19/England/BRIS-12F538/2020\|EPI_ISL_488240\|20 | ···················· | ···················· | ···················· | ···················· | ···················· | ···················· | |
| E | 6464 hCoV-19/England/BRIS-12F7C9/2020\|EPI_ISL_488241\|20 | ···················· | ···················· | ···················· | ···················· | ···················· | ···················· | |
| E | 6465 hCoV-19/England/BRIS-12E9C4/2020\|EPI_ISL_488243\|20 | ···················· | ····A··············· | ···················· | ···················· | ···················· | ···················· | |

| | | | | | | | | | |
|---|---|---|---|---|---|---|---|---|---|
| E | 6466 | hCoV-19/England/BRIS-12F5B0/2020\|EPI_ISL_488244\|20 | ···················· | ···················· | ···················· | ···················· | ···················· | ···················· | |
| E | 6467 | hCoV-19/England/BRIS-12EF77/2020\|EPI_ISL_488245\|20 | ···················· | ···················· | ···················· | ···················· | ···················· | ···················· | |
| E | 6468 | hCoV-19/England/BRIS-12E7CA/2020\|EPI_ISL_488246\|20 | ···················· | ···················· | ···················· | ···················· | ···················· | ···················· | |
| E | 6469 | hCoV-19/England/BRIS-12E87C/2020\|EPI_ISL_488247\|20 | ···················· | ···················· | ···················· | ···················· | ···················· | ···················· | |
| E | 6470 | hCoV-19/England/BRIS-12EF86/2020\|EPI_ISL_488248\|20 | ···················· | ···················· | ··T················· | ···················· | ···················· | ···················· | |
| E | 6471 | hCoV-19/England/BRIS-12EE10/2020\|EPI_ISL_488249\|20 | ···················· | ···················· | ···················· | ···················· | ···················· | ···················· | |
| E | 6472 | hCoV-19/England/BRIS-12F84E/2020\|EPI_ISL_488250\|20 | ···················· | ···················· | ···················· | ···················· | ···················· | ···················· | |
| E | 6473 | hCoV-19/England/BRIS-12FA48/2020\|EPI_ISL_488251\|20 | ···················· | ···················· | ···················· | ···················· | ···················· | ···················· | |
| E | 6474 | hCoV-19/England/BRIS-12ED13/2020\|EPI_ISL_488252\|20 | ···················· | ···················· | ···················· | ···················· | ···················· | ···················· | |
| E | 6475 | hCoV-19/England/BRIS-12F617/2020\|EPI_ISL_488253\|20 | ···················· | ···················· | ···················· | ···················· | ···················· | ···················· | |
| E | 6476 | hCoV-19/England/BRIS-12F6BD/2020\|EPI_ISL_488254\|20 | ···················· | ···················· | ···················· | ···················· | ···················· | ···················· | |
| E | 6477 | hCoV-19/England/BRIS-12E6CD/2020\|EPI_ISL_488255\|20 | ···················· | ···················· | ···················· | ···················· | ···················· | ···················· | |
| E | 6478 | hCoV-19/England/BRIS-12E812/2020\|EPI_ISL_488256\|20 | ···················· | ···················· | ···················· | ···················· | ···················· | ···················· | |
| E | 6479 | hCoV-19/England/BRIS-12ED31/2020\|EPI_ISL_488257\|20 | ···················· | ···················· | ···················· | ···················· | ···················· | ···················· | |
| E | 6480 | hCoV-19/Northern Ireland/NIRE-10010C/2020\|EPI_ISL_ | ···················· | ···················· | ···················· | ···················· | ···················· | ···················· | |
| E | 6481 | hCoV-19/Northern Ireland/NIRE-103479/2020\|EPI_ISL_ | ···················· | ···················· | ···················· | ···················· | ···················· | ···················· | |
| E | 6482 | hCoV-19/Northern Ireland/NIRE-10338B/2020\|EPI_ISL_ | ···················· | ···················· | ···················· | ···················· | ···················· | ···················· | |
| E | 6483 | hCoV-19/Northern Ireland/NIRE-FCD79/2020\|EPI_ISL_4 | ···················· | ···················· | ···················· | ···················· | ···················· | ···················· | |
| E | 6484 | hCoV-19/Northern Ireland/NIRE-FCDC4/2020\|EPI_ISL_4 | ···················· | ···················· | ···················· | ···················· | ···················· | ···················· | |
| E | 6485 | hCoV-19/Scotland/GCVR-171986/2020\|EPI_ISL_489380\|2 | ···················· | ···················· | ···················· | ···················· | ···················· | ···················· | |
| E | 6486 | hCoV-19/England/CAMB-1ACC6B/2020\|EPI_ISL_489381\|20 | ···················· | ···················· | ···················· | ···················· | ···················· | ···················· | |
| E | 6487 | hCoV-19/England/CAMB-1ACE1A/2020\|EPI_ISL_489382\|20 | ···················· | ···················· | ···················· | ···················· | ···················· | ···················· | |
| E | 6488 | hCoV-19/England/CAMB-1ACD0E/2020\|EPI_ISL_489383\|20 | ···················· | ···················· | ···················· | ···················· | ···················· | ···················· | |
| E | 6489 | hCoV-19/England/CAMB-1AC876/2020\|EPI_ISL_489384\|20 | ···················· | ···················· | ···················· | ···················· | ···················· | ···················· | |
| E | 6490 | hCoV-19/England/CAMB-1ACAE9/2020\|EPI_ISL_489386\|20 | ···················· | ···················· | ···················· | ···················· | ······T············· | ···················· | |
| E | 6491 | hCoV-19/England/CAMB-1AD5D8/2020\|EPI_ISL_489387\|20 | ···················· | ···················· | ···················· | ···················· | ···················· | ···················· | |
| E | 6492 | hCoV-19/England/CAMB-1AC524/2020\|EPI_ISL_489503\|20 | ···················· | ···················· | ···················· | ···················· | ···················· | ···················· | |
| E | 6493 | hCoV-19/England/CAMB-1AC8FE/2020\|EPI_ISL_489504\|20 | ···················· | ···················· | ···················· | ···················· | ···················· | ···················· | |
| E | 6494 | hCoV-19/England/CAMB-1ACE74/2020\|EPI_ISL_489505\|20 | ···················· | ···················· | ···················· | ···················· | ···················· | ···················· | |
| E | 6495 | hCoV-19/England/CAMB-1AC6F4/2020\|EPI_ISL_489506\|20 | ···················· | ···················· | ···················· | ···················· | ···················· | ···················· | |
| E | 6496 | hCoV-19/England/CAMB-1AC74C/2020\|EPI_ISL_489507\|20 | ···················· | ···················· | ···················· | ···················· | ···················· | ···················· | |
| E | 6497 | hCoV-19/England/CAMB-1ACC7A/2020\|EPI_ISL_489508\|20 | ···················· | ···················· | ···················· | ···················· | ···················· | ···················· | |
| E | 6498 | hCoV-19/England/CAMB-1AC973/2020\|EPI_ISL_489510\|20 | ···················· | ···················· | ···················· | ···················· | ···················· | ···················· | |
| E | 6499 | hCoV-19/England/CAMB-1AD63F/2020\|EPI_ISL_489511\|20 | ···················· | ···················· | ···················· | ···················· | ···················· | ···················· | |
| E | 6500 | hCoV-19/England/CAMB-1AC5BB/2020\|EPI_ISL_489512\|20 | ···················· | ···················· | ···················· | ···················· | ···················· | ···················· | |
| E | 6501 | hCoV-19/England/CAMB-1AC76A/2020\|EPI_ISL_489514\|20 | ···················· | ···················· | ···················· | ···················· | ···················· | ···················· | |
| E | 6502 | hCoV-19/England/CAMB-1AD67B/2020\|EPI_ISL_489517\|20 | ···················· | ···················· | ···················· | ···················· | ···················· | ···················· | |
| E | 6503 | hCoV-19/England/CAMB-1AC542/2020\|EPI_ISL_489518\|20 | ···················· | ···················· | ···················· | ···················· | ···················· | ···················· | |
| E | 6504 | hCoV-19/England/CAMB-1ACC4D/2020\|EPI_ISL_489519\|20 | ···················· | ···················· | ···················· | ···················· | ···················· | ···················· | |
| E | 6505 | hCoV-19/England/CAMB-1ACCF2/2020\|EPI_ISL_489520\|20 | ···················· | ·A·················· | ···················· | ···················· | ···················· | ···················· | |
| E | 6506 | hCoV-19/England/CAMB-1ACE92/2020\|EPI_ISL_489521\|20 | ···················· | ···················· | ···················· | ···················· | ···················· | ···················· | |
| E | 6507 | hCoV-19/England/CAMB-1ACF9F/2020\|EPI_ISL_489522\|20 | ···················· | ···················· | ···················· | ···················· | ···················· | ···················· | |
| E | 6508 | hCoV-19/England/CAMB-1AC937/2020\|EPI_ISL_489524\|20 | ···················· | ···················· | ···················· | ···················· | ···················· | ···················· | |
| E | 6509 | hCoV-19/England/CAMB-1ACDB3/2020\|EPI_ISL_489525\|20 | ···················· | ···················· | ···················· | ···················· | ···················· | ···················· | |
| E | 6510 | hCoV-19/England/CAMB-1ACDFF/2020\|EPI_ISL_489526\|20 | ···················· | ···················· | ···················· | ···················· | ···················· | ···················· | |
| E | 6511 | hCoV-19/England/CAMB-1AC481/2020\|EPI_ISL_489527\|20 | ···················· | ···················· | ···················· | ···················· | ···················· | ···················· | |
| E | 6512 | hCoV-19/England/CAMB-1ACEDE/2020\|EPI_ISL_489531\|20 | ···················· | ···················· | ···················· | ···················· | ···················· | ···················· | |
| E | 6513 | hCoV-19/England/CAMB-1AC867/2020\|EPI_ISL_489534\|20 | ···················· | ···················· | ···················· | ···················· | ···················· | ···················· | |
| E | 6514 | hCoV-19/England/CAMB-1AD59C/2020\|EPI_ISL_489535\|20 | ···················· | ···················· | ···················· | ···················· | ···················· | ···················· | |
| E | 6515 | hCoV-19/Scotland/GCVR-171995/2020\|EPI_ISL_489536\|2 | ···················· | ···················· | ···················· | ···················· | ···················· | ···················· | |
| E | 6516 | hCoV-19/Scotland/GCVR-171B08/2020\|EPI_ISL_489537\|2 | ···················· | ···················· | ···················· | ···················· | ···················· | ···················· | |
| E | 6517 | hCoV-19/England/CAMB-1ACC3E/2020\|EPI_ISL_489540\|20 | ···················· | ···················· | ···················· | ···················· | ···················· | ···················· | |
| E | 6518 | hCoV-19/England/CAMB-1ACA8F/2020\|EPI_ISL_489541\|20 | ···················· | ···················· | ···················· | ···················· | ···················· | ···················· | |
| E | 6519 | hCoV-19/Scotland/GCVR-171A47/2020\|EPI_ISL_489543\|2 | ···················· | ···················· | ···················· | ···················· | ···················· | ···················· | |
| E | 6520 | hCoV-19/England/CAMB-1ACF26/2020\|EPI_ISL_489544\|20 | ···················· | ·A·················· | ···················· | ···················· | ···················· | ···················· | |
| E | 6521 | hCoV-19/England/CAMB-1AC6C7/2020\|EPI_ISL_489545\|20 | ···················· | ···················· | ···················· | ···················· | ···················· | ···················· | |
| E | 6522 | hCoV-19/England/CAMB-1AC7E2/2020\|EPI_ISL_489546\|20 | ···················· | ···················· | ···················· | ···················· | ···················· | ···················· | |
| E | 6523 | hCoV-19/Scotland/GCVR-171A0B/2020\|EPI_ISL_489547\|2 | ···················· | ···················· | ···················· | ···················· | ···················· | ···················· | |
| E | 6524 | hCoV-19/England/CAMB-1AC928/2020\|EPI_ISL_489549\|20 | ···················· | ···················· | ···················· | ···················· | ···················· | ···················· | |
| E | 6525 | hCoV-19/England/CAMB-1AC90A/2020\|EPI_ISL_489550\|20 | ···················· | ···················· | ···················· | ···················· | ···················· | ···················· | |
| E | 6526 | hCoV-19/England/CAMB-1AC9EC/2020\|EPI_ISL_489552\|20 | ···················· | ···················· | ···················· | ···················· | ···················· | ···················· | |
| E | 6527 | hCoV-19/England/CAMB-1AD374/2020\|EPI_ISL_489553\|20 | ···················· | ···················· | ···················· | ···················· | ···················· | ···················· | |
| E | 6528 | hCoV-19/England/CAMB-1ACBAA/2020\|EPI_ISL_489554\|20 | ···················· | ···················· | ···················· | ···················· | ···················· | ···················· | |
| E | 6529 | hCoV-19/England/CAMB-1AC9DD/2020\|EPI_ISL_489557\|20 | ···················· | ···················· | ···················· | ···················· | ···················· | ···················· | |
| E | 6530 | hCoV-19/England/CAMB-1ACF08/2020\|EPI_ISL_489558\|20 | ···················· | ···················· | ···················· | ···················· | ···················· | ···················· | |
| E | 6531 | hCoV-19/England/CAMB-1AC612/2020\|EPI_ISL_489559\|20 | ···················· | ···················· | ···················· | ···················· | ···················· | ···················· | |
| E | 6532 | hCoV-19/England/CAMB-1AC64F/2020\|EPI_ISL_489560\|20 | ···················· | ···················· | ···················· | ···················· | ···················· | ···················· | |
| E | 6533 | hCoV-19/England/CAMB-1AC72E/2020\|EPI_ISL_489562\|20 | ···················· | ···················· | ···················· | ···················· | ···················· | ···················· | |
| E | 6534 | hCoV-19/England/CAMB-1AD4AE/2020\|EPI_ISL_489563\|20 | ···················· | ···················· | ···················· | ···················· | ···················· | ···················· | |
| E | 6535 | hCoV-19/England/CAMB-1ACB40/2020\|EPI_ISL_489564\|20 | ···················· | ···················· | ···················· | ···················· | ···················· | ···················· | |
| E | 6536 | hCoV-19/England/CAMB-1AD49F/2020\|EPI_ISL_489565\|20 | ···················· | ···················· | ···················· | ···················· | ···················· | ···················· | |
| E | 6537 | hCoV-19/England/CAMB-1AC67C/2020\|EPI_ISL_489568\|20 | ···················· | ···················· | ···················· | ···················· | ···················· | ···················· | |
| E | 6538 | hCoV-19/England/CAMB-1AC490/2020\|EPI_ISL_489569\|20 | ···················· | ···················· | ···················· | ···················· | ···················· | ···················· | |
| E | 6539 | hCoV-19/England/CAMB-1AC82B/2020\|EPI_ISL_489570\|20 | ···················· | ···················· | ···················· | ···················· | ···················· | ···················· | |
| E | 6540 | hCoV-19/England/CAMB-1AC946/2020\|EPI_ISL_489571\|20 | ···················· | ···················· | ···················· | ···················· | ·················C·· | ···················· | 1 |
| E | 6541 | hCoV-19/England/CAMB-1AC919/2020\|EPI_ISL_489572\|20 | ···················· | ···················· | ···················· | ···················· | ···················· | ···················· | |
| E | 6542 | hCoV-19/England/CAMB-1AC71F/2020\|EPI_ISL_489573\|20 | ···················· | ···················· | ···················· | ···················· | ···················· | ···················· | |
| E | 6543 | hCoV-19/England/CAMB-1ACF80/2020\|EPI_ISL_489575\|20 | ···················· | ···················· | ···················· | ···················· | ···················· | ···················· | |
| E | 6544 | hCoV-19/England/CAMB-1AC7F1/2020\|EPI_ISL_489576\|20 | ···················· | ···················· | ···················· | ···················· | ···················· | ···················· | |
| E | 6545 | hCoV-19/England/CAMB-1AC991/2020\|EPI_ISL_489577\|20 | ···················· | ···················· | ···················· | ···················· | ···················· | ···················· | |

| | | | | | | | | |
|---|---|---|---|---|---|---|---|---|
| E | 6546 hCoV-19/Scotland/GCVR-170820/2020\|EPI_ISL_489578\|2 | ···················· | ···················· | ···················· | ···················· | ···················· | ···················· | |
| E | 6547 hCoV-19/Scotland/GCVR-16FB49/2020\|EPI_ISL_489579\|2 | ···················· | ···················· | ···················· | ···················· | ···················· | ···················· | |
| E | 6548 hCoV-19/Scotland/GCVR-171810/2020\|EPI_ISL_489580\|2 | ···················· | ···················· | ···················· | ···················· | ···················· | ···················· | |
| E | 6549 hCoV-19/Scotland/GCVR-16FEB9/2020\|EPI_ISL_489584\|2 | ···················· | ···················· | ···················· | ···················· | ···················· | ···················· | |
| E | 6550 hCoV-19/Scotland/GCVR-16FB67/2020\|EPI_ISL_489585\|2 | ···················· | ···················· | ···················· | ···················· | ···················· | ···················· | |
| E | 6551 hCoV-19/Scotland/GCVR-1717F5/2020\|EPI_ISL_489586\|2 | ···················· | ···················· | ···················· | ···················· | ···················· | ···················· | |
| E | 6552 hCoV-19/Scotland/GCVR-17083F/2020\|EPI_ISL_489587\|2 | ···················· | ···················· | ···················· | ···················· | ···················· | ···················· | |
| E | 6553 hCoV-19/Scotland/GCVR-16FE8C/2020\|EPI_ISL_489588\|2 | ···················· | ···················· | ···················· | ···················· | ···················· | ···················· | |
| E | 6554 hCoV-19/England/LIVE-A1B5B/2020\|EPI_ISL_490354\|202 | ···················· | ···················· | ···················· | ···················· | ···················· | ···················· | |
| E | 6555 hCoV-19/England/LIVE-A1C0D/2020\|EPI_ISL_490359\|202 | ···················· | ···················· | ···················· | ···················· | ···················· | ···················· | |
| E | 6556 hCoV-19/England/LIVE-A1D19/2020\|EPI_ISL_490364\|202 | ···················· | ···················· | ···················· | ···················· | ···················· | ···················· | |
| E | 6557 hCoV-19/England/LIVE-A1D28/2020\|EPI_ISL_490365\|202 | ···················· | ···················· | ···················· | ···················· | ···················· | ···················· | |
| E | 6558 hCoV-19/England/LIVE-A1F9B/2020\|EPI_ISL_490370\|202 | ···················· | ···················· | ···················· | ···················· | ···················· | ···················· | |
| E | 6568 hCoV-19/England/NORW-EE24E/2020\|EPI_ISL_490519\|202 | ···················· | ···················· | ···················· | ···················· | ···················· | ···················· | |
| E | 6569 hCoV-19/England/NORW-EE26C/2020\|EPI_ISL_490520\|202 | ···················· | ···················· | ···················· | ···················· | NNN················· | ···················· | 1 |
| E | 6572 hCoV-19/England/NORW-EE2E4/2020\|EPI_ISL_490528\|202 | ···················· | ···················· | ···················· | ···················· | NNN················· | ···················· | 1 |
| E | 6573 hCoV-19/England/NORW-EE30F/2020\|EPI_ISL_490529\|202 | ···················· | ···················· | ···················· | ···················· | NN·················· | ···················· | |
| E | 6579 hCoV-19/England/NORW-EE41B/2020\|EPI_ISL_490540\|202 | ···················· | ···················· | ···················· | ···················· | ···················· | ···················· | |
| E | 6580 hCoV-19/England/NORW-EE42A/2020\|EPI_ISL_490541\|202 | ···················· | ···················· | ···················· | ···················· | ···················· | ···················· | |
| E | 6588 hCoV-19/England/NOTT-112662/2020\|EPI_ISL_490561\|20 | ···················· | ···················· | ···················· | ···················· | ···················· | ···················· | |
| E | 6589 hCoV-19/England/NOTT-11269F/2020\|EPI_ISL_490562\|20 | ···················· | ···················· | ···················· | ···················· | ···················· | ···················· | |
| E | 6590 hCoV-19/England/NOTT-1126AE/2020\|EPI_ISL_490563\|20 | ···················· | ···················· | ···················· | ···················· | ···················· | ···················· | |
| E | 6591 hCoV-19/England/NOTT-1126DB/2020\|EPI_ISL_490566\|20 | ···················· | ···················· | ···················· | ···················· | ···················· | ···················· | |
| E | 6592 hCoV-19/England/NOTT-1126EA/2020\|EPI_ISL_490567\|20 | ···················· | ···················· | ···················· | ···················· | ···················· | ···················· | |
| E | 6593 hCoV-19/England/NOTT-1126F9/2020\|EPI_ISL_490568\|20 | ···················· | ···················· | ···················· | ···················· | ···················· | ···················· | |
| E | 6594 hCoV-19/England/NOTT-112705/2020\|EPI_ISL_490569\|20 | ···················· | ···················· | ···················· | ···················· | ···················· | ···················· | |
| E | 6595 hCoV-19/Scotland/EDB3595/2020\|EPI_ISL_491680\|2020- | ···················· | ·A·················· | ···················· | ···················· | ···················· | ···················· | |
| E | 6596 hCoV-19/Scotland/EDB2019/2020\|EPI_ISL_491681\|2020- | ···················· | ···················· | ···················· | ···················· | ···················· | ···················· | |
| E | 6597 hCoV-19/Scotland/EDB2262/2020\|EPI_ISL_491682\|2020- | ···················· | ···················· | ···················· | ···················· | ···················· | ···················· | |
| E | 6598 hCoV-19/Scotland/EDB2255/2020\|EPI_ISL_491685\|2020- | ···················· | ···················· | ···················· | ···················· | ···················· | ···················· | |
| E | 6599 hCoV-19/Scotland/EDB2000/2020\|EPI_ISL_491686\|2020- | ···················· | ···················· | ···················· | ···················· | ···················· | ···················· | |
| E | 6600 hCoV-19/Scotland/EDB1834/2020\|EPI_ISL_491687\|2020- | ···················· | ···················· | ···················· | ···················· | ···················· | ···················· | |
| E | 6601 hCoV-19/Scotland/EDB404/2020\|EPI_ISL_491689\|2020-0 | ···················· | ···················· | ···················· | ···················· | ···················· | ···················· | |
| E | 6602 hCoV-19/Scotland/EDB1853/2020\|EPI_ISL_491692\|2020- | ···················· | ···················· | ···················· | ···················· | ···················· | ···················· | |
| E | 6603 hCoV-19/Scotland/EDB2138/2020\|EPI_ISL_491693\|2020- | ···················· | ···················· | ···················· | ···················· | ···················· | ···················· | |
| E | 6604 hCoV-19/Scotland/EDB2248/2020\|EPI_ISL_491695\|2020- | ···················· | ·A·················· | ···················· | ···················· | ···················· | ···················· | |
| E | 6605 hCoV-19/Scotland/EDB6741/2020\|EPI_ISL_491696\|2020- | ···················· | ·A·················· | ···················· | ···················· | ···················· | ···················· | |
| E | 6606 hCoV-19/Scotland/EDB1472/2020\|EPI_ISL_491698\|2020- | ···················· | ···················· | ···················· | ···················· | ···················· | ···················· | |
| E | 6607 hCoV-19/Scotland/EDB1342/2020\|EPI_ISL_491699\|2020- | ···················· | ·A·················· | ···················· | ···················· | ···················· | ···················· | |
| E | 6608 hCoV-19/Wales/PHWC-1668E1/2020\|EPI_ISL_494057\|2020 | ···················· | ···················· | ···················· | ···················· | ···················· | ···················· | |
| E | 6609 hCoV-19/Wales/PHWC-1668F0/2020\|EPI_ISL_494058\|2020 | ···················· | ···················· | ···················· | ···················· | ···················· | ···················· | |
| E | 6610 hCoV-19/Wales/PHWC-16690C/2020\|EPI_ISL_494059\|2020 | ···················· | ···················· | ···················· | ···················· | ···················· | ···················· | |
| E | 6611 hCoV-19/Wales/PHWC-16691B/2020\|EPI_ISL_494060\|2020 | ···················· | ···················· | ···················· | ···················· | ···················· | ···················· | |
| E | 6613 hCoV-19/Wales/PHWC-166957/2020\|EPI_ISL_494062\|2020 | ···················· | ···················· | ···················· | ···················· | ···················· | ···················· | |
| E | 6614 hCoV-19/Wales/PHWC-166966/2020\|EPI_ISL_494063\|2020 | ···················· | ···················· | ···················· | ···················· | ···················· | ···················· | |
| E | 6616 hCoV-19/Wales/PHWC-166984/2020\|EPI_ISL_494065\|2020 | ···················· | ···················· | ···················· | ···················· | ···················· | ···················· | |
| E | 6617 hCoV-19/Wales/PHWC-1669B1/2020\|EPI_ISL_494066\|2020 | ···················· | ···················· | ···················· | ···················· | ···················· | ···················· | |
| E | 6618 hCoV-19/Wales/PHWC-1669EE/2020\|EPI_ISL_494068\|2020 | ···················· | ···················· | ···················· | ···················· | ···················· | ···················· | |
| E | 6619 hCoV-19/Wales/PHWC-166A09/2020\|EPI_ISL_494069\|2020 | ···················· | ···················· | ···················· | ···················· | ·NN················· | ···················· | |
| E | 6620 hCoV-19/Wales/PHWC-166A18/2020\|EPI_ISL_494070\|2020 | ···················· | ···················· | ···················· | ···················· | ···················· | ···················· | |
| E | 6621 hCoV-19/Wales/PHWC-166A36/2020\|EPI_ISL_494071\|2020 | ···················· | ···················· | ···················· | ···················· | ···················· | ···················· | |
| E | 6622 hCoV-19/Wales/PHWC-166A54/2020\|EPI_ISL_494073\|2020 | ···················· | ···················· | ···················· | ···················· | ···················· | ···················· | |
| E | 6623 hCoV-19/Wales/PHWC-166A63/2020\|EPI_ISL_494074\|2020 | ···················· | ···················· | ···················· | ···················· | ···················· | ···················· | |
| E | 6624 hCoV-19/Wales/PHWC-166A72/2020\|EPI_ISL_494075\|2020 | ···················· | ···················· | ···················· | ···················· | ···················· | ···················· | |
| E | 6625 hCoV-19/Wales/PHWC-166A81/2020\|EPI_ISL_494076\|2020 | ···················· | ···················· | ···················· | ···················· | ···················· | ···················· | |
| E | 6626 hCoV-19/Wales/PHWC-166ABE/2020\|EPI_ISL_494077\|2020 | ···················· | ···················· | ···················· | ···················· | ···················· | ···················· | |
| E | 6627 hCoV-19/Wales/PHWC-166BAC/2020\|EPI_ISL_494089\|2020 | ···················· | ···················· | ···················· | ···················· | ···················· | ···················· | |
| E | 6628 hCoV-19/Wales/PHWC-166BBB/2020\|EPI_ISL_494090\|2020 | ···················· | ···················· | ···················· | ···················· | ···················· | ···················· | |
| E | 6629 hCoV-19/Wales/PHWC-166BCA/2020\|EPI_ISL_494091\|2020 | ···················· | ···················· | ···················· | ···················· | ···················· | ···················· | |
| E | 6631 hCoV-19/Wales/PHWC-166BF7/2020\|EPI_ISL_494093\|2020 | ···················· | ···················· | ···················· | ···················· | ···················· | ···················· | |
| E | 6632 hCoV-19/Wales/PHWC-166C03/2020\|EPI_ISL_494094\|2020 | ···················· | ···················· | ···················· | ···················· | ···················· | ···················· | |
| E | 6636 hCoV-19/Wales/PHWC-166C8B/2020\|EPI_ISL_494099\|2020 | ···················· | ···················· | ···················· | ···················· | ···················· | ···················· | |
| E | 6637 hCoV-19/USA/FL-BPHL-0481/2020\|EPI_ISL_495387\|2020- | ···················· | ···················· | ···················· | ···················· | ···················· | ···················· | |
| E | 6639 hCoV-19/USA/FL-BPHL-0483/2020\|EPI_ISL_495389\|2020- | ···················· | ···················· | ···················· | ···················· | ···················· | ···················· | |
| E | 6640 hCoV-19/USA/FL-BPHL-0484/2020\|EPI_ISL_495390\|2020- | ···················· | ···················· | ···················· | ···················· | ···················· | ···················· | |
| E | 6641 hCoV-19/USA/FL-BPHL-0487/2020\|EPI_ISL_495393\|2020- | ···················· | ···················· | ···················· | ···················· | ···················· | ···················· | |
| E | 6643 hCoV-19/USA/FL-BPHL-0489/2020\|EPI_ISL_495395\|2020- | ···················· | ···················· | ···················· | ···················· | ···················· | ···················· | |
| E | 6644 hCoV-19/USA/FL-BPHL-0490/2020\|EPI_ISL_495396\|2020- | ···················· | ···················· | ···················· | ···················· | ···················· | ···················· | |
| E | 6645 hCoV-19/USA/FL-BPHL-0491/2020\|EPI_ISL_495397\|2020- | ···················· | ···················· | ···················· | ···················· | ··················NN | ···················· | 1 |
| E | 6647 hCoV-19/USA/FL-BPHL-0493/2020\|EPI_ISL_495399\|2020- | ···················· | ···················· | ···················· | ···················· | ···················· | ···················· | |
| E | 6648 hCoV-19/lion/USA/NY-2-041520/2020\|EPI_ISL_566036\|2 | ···················· | ···················· | ···················· | ···················· | ···················· | ···················· | |
| E | 6649 hCoV-19/lion/USA/NY-3-041520/2020\|EPI_ISL_566037\|2 | ···················· | ···················· | ···················· | ···················· | ···················· | ···················· | |
| E | 6650 hCoV-19/lion/USA/NY-041520/2020\|EPI_ISL_566038\|202 | ···················· | ···················· | ···················· | ···················· | ···················· | ···················· | |
| E | 6651 hCoV-19/tiger/USA/NY-2-040420/2020\|EPI_ISL_566039\| | ···················· | ···················· | ···················· | ···················· | ···················· | ···················· | |
| E | 6652 hCoV-19/tiger/USA/NY-P3/2020\|EPI_ISL_566040\|2020-0 | ···················· | ···················· | ···················· | ···················· | ··C················· | ···················· | |
| E | 6653 hCoV-19/tiger/USA/NY-3-040420/2020\|EPI_ISL_566041\| | ···················· | ···················· | ···················· | ···················· | ···················· | ···················· | |
| E | 6654 hCoV-19/tiger/USA/NY-4-040420/2020\|EPI_ISL_566042\| | ···················· | ···················· | ···················· | ···················· | ···················· | ···················· | |
| E | 6655 hCoV-19/tiger/USA/NY-P4/2020\|EPI_ISL_566043\|2020-0 | ···················· | ···················· | ···················· | ···················· | ···················· | ···················· | |
| E | 6656 hCoV-19/lion/USA/NY-2/2020\|EPI_ISL_566044\|2020-04- | ···················· | ···················· | ···················· | ···················· | ···················· | ···················· | |
| E | 6657 hCoV-19/England/BRIS-12E092/2020\|EPI_ISL_482000\|20 | ···················· | ···················· | ···················· | ···················· | ···················· | ···················· | |

| | | | | | | | |
|---|---|---|---|---|---|---|---|
| E | 6658 hCoV-19/England/BRIS-12DBFC/2020\|EPI_ISL_482001\|20 | ···················· | ···················· | ···················· | ···················· | ···················· | ···················· |
| E | 6659 hCoV-19/England/BRIS-12DE5D/2020\|EPI_ISL_482002\|20 | ···················· | ···················· | ···················· | ···················· | ···················· | ···················· |
| E | 6660 hCoV-19/England/BRIS-12E108/2020\|EPI_ISL_482003\|20 | ···················· | ···A················ | ···················· | ···················· | ···················· | ···················· |
| E | 6661 hCoV-19/England/BRIS-12E126/2020\|EPI_ISL_482004\|20 | ···················· | ···················· | ···················· | ···················· | ···················· | ···················· |
| E | 6662 hCoV-19/England/BRIS-12DE8A/2020\|EPI_ISL_482005\|20 | ···················· | ···················· | ···················· | ···················· | ···················· | ···················· |
| E | 6663 hCoV-19/England/BRIS-12DC53/2020\|EPI_ISL_482007\|20 | ···················· | ···················· | ···················· | ···················· | ···················· | ···················· |
| E | 6664 hCoV-19/England/BRIS-12E250/2020\|EPI_ISL_482009\|20 | ···················· | ···················· | ···················· | ···················· | ···················· | ···················· |
| E | 6665 hCoV-19/England/BRIS-12DF3C/2020\|EPI_ISL_482010\|20 | ···················· | ···················· | ···················· | ···················· | ···················· | ···················· |
| E | 6666 hCoV-19/England/BRIS-12DE02/2020\|EPI_ISL_482011\|20 | ···················· | ···················· | ···················· | ···················· | ···················· | ···················· |
| E | 6667 hCoV-19/England/BRIS-12E205/2020\|EPI_ISL_482013\|20 | ···················· | ···················· | ···················· | ···················· | ···················· | ···················· |
| E | 6668 hCoV-19/England/BRIS-12DE6C/2020\|EPI_ISL_482015\|20 | ···················· | ···················· | ···················· | ···················· | ···················· | ···················· |
| E | 6669 hCoV-19/England/BRIS-12E0CF/2020\|EPI_ISL_482017\|20 | ···················· | ···················· | ···················· | ···················· | ···················· | ···················· |
| E | 6670 hCoV-19/England/BRIS-12DF78/2020\|EPI_ISL_482018\|20 | ···················· | ···················· | ···················· | ···················· | ···················· | ···················· |
| E | 6671 hCoV-19/England/BRIS-12E1EA/2020\|EPI_ISL_482020\|20 | ···················· | ···················· | ···················· | ···················· | ···················· | ···················· |
| E | 6672 hCoV-19/England/BRIS-12E0A1/2020\|EPI_ISL_482021\|20 | ···················· | ···················· | ···················· | ···················· | ···················· | ···················· |
| E | 6673 hCoV-19/Northern Ireland/NIRE-104214/2020\|EPI_ISL_ | ···················· | ···················· | ···················· | ···················· | ···················· | ···················· |
| E | 6674 hCoV-19/Northern Ireland/NIRE-FF56F/2020\|EPI_ISL_4 | ···················· | ···················· | ···················· | ···················· | ···················· | ···················· |
| E | 6675 hCoV-19/Northern Ireland/NIRE-103E7B/2020\|EPI_ISL_ | ···················· | ···················· | ···················· | ···················· | ···················· | ···················· |
| E | 6676 hCoV-19/Northern Ireland/NIRE-103F69/2020\|EPI_ISL_ | ···················· | ···················· | ···················· | ···················· | ···················· | ···················· |
| E | 6677 hCoV-19/Northern Ireland/NIRE-10429C/2020\|EPI_ISL_ | ···················· | ···················· | ···················· | ···················· | ···················· | ···················· |
| E | 6678 hCoV-19/Northern Ireland/NIRE-FF58D/2020\|EPI_ISL_4 | ···················· | ···················· | ···················· | ···················· | ···················· | ···················· |
| E | 6679 hCoV-19/Northern Ireland/NIRE-103F87/2020\|EPI_ISL_ | ···················· | ···················· | ···················· | ···················· | ···················· | ···················· |
| E | 6680 hCoV-19/Northern Ireland/NIRE-103FA5/2020\|EPI_ISL_ | ···················· | ···················· | ···················· | ···················· | ···················· | ···················· |
| E | 6681 hCoV-19/Northern Ireland/NIRE-103D14/2020\|EPI_ISL_ | ···················· | ···················· | ···················· | ···················· | ···················· | ···················· |
| E | 6682 hCoV-19/Northern Ireland/NIRE-103E99/2020\|EPI_ISL_ | ···················· | ···················· | ···················· | ···················· | ···················· | ···················· |
| E | 6683 hCoV-19/Northern Ireland/NIRE-103EC6/2020\|EPI_ISL_ | ···················· | ···················· | ···················· | ···················· | ···················· | ···················· |
| E | 6684 hCoV-19/Northern Ireland/NIRE-FF259/2020\|EPI_ISL_4 | ···················· | ···················· | ···················· | ···················· | ···················· | ···················· |
| E | 6685 hCoV-19/Northern Ireland/NIRE-FF268/2020\|EPI_ISL_4 | ···················· | ···················· | ···················· | ···················· | ···················· | ···················· |
| E | 6686 hCoV-19/Northern Ireland/NIRE-103EB7/2020\|EPI_ISL_ | ···················· | ···················· | ···················· | ···················· | ···················· | ···················· |
| E | 6687 hCoV-19/England/TBSD-2488FEB/2020\|EPI_ISL_482058\|2 | ···················· | ···················· | ···················· | ···················· | ···················· | ···················· |
| E | 6688 hCoV-19/England/TBSD-2488DC3/2020\|EPI_ISL_482060\|2 | ···················· | ···················· | ···················· | ···················· | ···················· | ···················· |
| E | 6689 hCoV-19/England/TBSD-2488E2A/2020\|EPI_ISL_482061\|2 | ···················· | ···A················ | ···················· | ···················· | ···················· | ···················· |
| E | 6690 hCoV-19/England/TBSD-2488F54/2020\|EPI_ISL_482062\|2 | ···················· | ···················· | ···················· | ···················· | ···················· | ···················· |
| E | 6691 hCoV-19/England/TBSD-2488E84/2020\|EPI_ISL_482064\|2 | ···················· | ···················· | ···················· | ···················· | ···················· | ···················· |
| E | 6692 hCoV-19/England/TBSD-248909C/2020\|EPI_ISL_482065\|2 | ···················· | ···················· | ···················· | ···················· | ···················· | ···················· |
| E | 6693 hCoV-19/England/TBSD-2488E48/2020\|EPI_ISL_482067\|2 | ···················· | ···················· | ···················· | ···················· | ···················· | ···················· |
| E | 6694 hCoV-19/England/TBSD-2488F18/2020\|EPI_ISL_482068\|2 | ···················· | ···M················ | ···················· | ···················· | ···················· | ···················· |
| E | 6695 hCoV-19/England/HECH-2599D55/2020\|EPI_ISL_482069\|2 | ···················· | ···················· | ···················· | ···················· | ···················· | ···················· |
| E | 6696 hCoV-19/Northern Ireland/NIRE-1041CC/2020\|EPI_ISL_ | ···················· | ···················· | ···················· | ···················· | ···················· | ···················· |
| E | 6697 hCoV-19/England/HECH-259A079/2020\|EPI_ISL_482071\|2 | ···················· | ···················· | ···················· | ···················· | ···················· | ···················· |
| E | 6698 hCoV-19/England/HECH-2599FC8/2020\|EPI_ISL_482072\|2 | ···················· | ···················· | ···················· | ···················· | ···················· | ···················· |
| E | 6699 hCoV-19/England/HECH-2599DEC/2020\|EPI_ISL_482073\|2 | ···················· | ···················· | ···················· | ···················· | ···················· | ···················· |
| E | 6700 hCoV-19/England/HECH-2599FAA/2020\|EPI_ISL_482074\|2 | ···················· | ···················· | ···················· | ···················· | ···················· | ···················· |
| E | 6701 hCoV-19/England/HECH-2599E9E/2020\|EPI_ISL_482076\|2 | ···················· | ···················· | ···················· | ···················· | ···················· | ···················· |
| E | 6702 hCoV-19/USA/CA-ALSR-0715-SAN/2020\|EPI_ISL_483320\|2 | ···················· | ···················· | ···················· | ···················· | ···················· | ···················· |
| E | 6703 hCoV-19/USA/CA-ALSR-0718-SAN/2020\|EPI_ISL_483321\|2 | ···················· | ···················· | ···················· | ···················· | ···················· | ···················· |
| E | 6704 hCoV-19/USA/CA-ALSR-0719-SAN/2020\|EPI_ISL_483322\|2 | ···················· | ···················· | ···················· | ···················· | ···················· | ···················· |
| E | 6705 hCoV-19/USA/CA-ALSR-0721-SAN/2020\|EPI_ISL_483323\|2 | ···················· | ···················· | ···················· | ···················· | ···················· | ···················· |
| E | 6706 hCoV-19/USA/CA-ALSR-0722-SAN/2020\|EPI_ISL_483324\|2 | ···················· | ·A·················· | ···················· | ···················· | ···················· | ···················· |
| E | 6707 hCoV-19/USA/CA-ALSR-0724-IPL/2020\|EPI_ISL_483325\|2 | ···················· | ·A·················· | ···················· | ···················· | ···················· | ···················· |
| E | 6708 hCoV-19/USA/CA-ALSR-0725-IPL/2020\|EPI_ISL_483326\|2 | ···················· | ···················· | ···················· | ···················· | ···················· | ···················· |
| E | 6709 hCoV-19/USA/CA-ALSR-0726-IPL/2020\|EPI_ISL_483327\|2 | ···················· | ·A·················· | ···················· | ···················· | ···················· | ···················· |
| E | 6710 hCoV-19/USA/CA-ALSR-0728-IPL/2020\|EPI_ISL_483328\|2 | ···················· | ·A·················· | ···················· | ···················· | ···················· | ···················· |
| E | 6711 hCoV-19/USA/CA-ALSR-0729-IPL/2020\|EPI_ISL_483329\|2 | ···················· | ·A·················· | ···················· | ···················· | ···················· | ···················· |
| E | 6712 hCoV-19/USA/CA-ALSR-0732-SAN/2020\|EPI_ISL_483330\|2 | ···················· | ·A·················· | ···················· | ···················· | ···················· | ···················· |
| E | 6713 hCoV-19/USA/CA-ALSR-0746-IPL/2020\|EPI_ISL_483331\|2 | ···················· | ···················· | ···················· | ···················· | ···················· | ···················· |
| E | 6714 hCoV-19/USA/CA-ALSR-0752-IPL/2020\|EPI_ISL_483332\|2 | ···················· | ·A·················· | ···················· | ···················· | ···················· | ···················· |
| E | 6715 hCoV-19/USA/CA-ALSR-0753-IPL/2020\|EPI_ISL_483333\|2 | ···················· | ·A·················· | ···················· | ···················· | ···················· | ···················· |
| E | 6716 hCoV-19/USA/CA-ALSR-0754-IPL/2020\|EPI_ISL_483334\|2 | ···················· | ·A·················· | ···················· | ···················· | ···················· | ···················· |
| E | 6717 hCoV-19/USA/CA-ALSR-0755-IPL/2020\|EPI_ISL_483335\|2 | ···················· | ···················· | ···················· | ···················· | ···················· | ···················· |
| E | 6718 hCoV-19/USA/CA-ALSR-0756-IPL/2020\|EPI_ISL_483336\|2 | ···················· | ···················· | ···················· | ···················· | ···················· | ···················· |
| E | 6719 hCoV-19/USA/CA-ALSR-0757-IPL/2020\|EPI_ISL_483337\|2 | ···················· | ·A·················· | ···················· | ···················· | ···················· | ···················· |
| E | 6720 hCoV-19/USA/CA-ALSR-0759-IPL/2020\|EPI_ISL_483338\|2 | ···················· | ·A·················· | ···················· | ···················· | ···················· | ···················· |
| E | 6721 hCoV-19/USA/CA-ALSR-0760-IPL/2020\|EPI_ISL_483339\|2 | ···················· | ·A·················· | ···················· | ···················· | ···················· | ···················· |
| E | 6722 hCoV-19/USA/CA-ALSR-0761-IPL/2020\|EPI_ISL_483340\|2 | ···················· | ···················· | ···················· | ···················· | ···················· | ···················· |
| E | 6723 hCoV-19/USA/CA-ALSR-0762-SAN/2020\|EPI_ISL_483341\|2 | ···················· | ···················· | ···················· | ···················· | ···················· | ···················· |
| E | 6724 hCoV-19/USA/CA-ALSR-0763-IPL/2020\|EPI_ISL_483342\|2 | ···················· | ···················· | ···················· | ···················· | ···················· | ···················· |
| E | 6725 hCoV-19/USA/CA-ALSR-0764-SAN/2020\|EPI_ISL_483343\|2 | ···················· | ···················· | ···················· | ···················· | ···················· | ···················· |
| E | 6726 hCoV-19/USA/CA-ALSR-0765-IPL/2020\|EPI_ISL_483344\|2 | ···················· | ···················· | ···················· | ···················· | ···················· | ···················· |
| E | 6727 hCoV-19/USA/CA-ALSR-0766-SAN/2020\|EPI_ISL_483345\|2 | ···················· | ···················· | ···················· | ···················· | ···················· | ···················· |
| E | 6728 hCoV-19/USA/CA-ALSR-0767-IPL/2020\|EPI_ISL_483346\|2 | ···················· | ·A·················· | ···················· | ···················· | ···················· | ···················· |
| E | 6729 hCoV-19/USA/CA-ALSR-0771-IPL/2020\|EPI_ISL_483347\|2 | ···················· | ·A·················· | ···················· | ···················· | ···················· | ···················· |
| E | 6730 hCoV-19/USA/CA-ALSR-0772-SAN/2020\|EPI_ISL_483348\|2 | ···················· | ···················· | ···················· | ···················· | ···················· | ···················· |
| E | 6731 hCoV-19/USA/CA-ALSR-0773-SAN/2020\|EPI_ISL_483349\|2 | ···················· | ·A·················· | ···················· | ···················· | ···················· | ···················· |
| E | 6732 hCoV-19/USA/CA-ALSR-0774-SAN/2020\|EPI_ISL_483350\|2 | ···················· | ···················· | ···················· | ···················· | ···················· | ···················· |
| E | 6733 hCoV-19/USA/CA-ALSR-0776-SAN/2020\|EPI_ISL_483351\|2 | ···················· | ···················· | ···················· | ···················· | ···················· | ···················· |
| E | 6734 hCoV-19/USA/CA-ALSR-0778-IPL/2020\|EPI_ISL_483352\|2 | ···················· | ·A·················· | ···················· | ···················· | ···················· | ···················· |
| E | 6735 hCoV-19/USA/CA-ALSR-0799-SAN/2020\|EPI_ISL_483366\|2 | ···················· | ···················· | ···················· | ···················· | ···················· | ···················· |
| E | 6736 hCoV-19/USA/CA-ALSR-0801-SAN/2020\|EPI_ISL_483368\|2 | ···················· | ···················· | ···················· | ···················· | ···················· | ···················· |
| E | 6737 hCoV-19/USA/CA-ALSR-0803-SAN/2020\|EPI_ISL_483370\|2 | ···················· | ···················· | ···················· | ···················· | ···················· | ···················· |

| | | | | | | | | | |
|---|---|---|---|---|---|---|---|---|---|
| E | 6738 | hCoV-19/USA/CA-ALSR-0805-SAN/2020\|EPI_ISL_483371\|2 | .................... | .................... | .................... | .................... | .................... | .................... | |
| E | 6739 | hCoV-19/USA/CA-ALSR-0806-SAN/2020\|EPI_ISL_483372\|2 | .................... | .................... | .................... | .................... | .................... | .................... | |
| E | 6740 | hCoV-19/USA/CA-ALSR-0809-SAN/2020\|EPI_ISL_483373\|2 | .................... | .................... | .................... | .................... | .................... | .................... | |
| E | 6741 | hCoV-19/USA/CA-ALSR-0810-SAN/2020\|EPI_ISL_483374\|2 | .................... | .................... | .................... | .................... | .................... | .................... | |
| E | 6742 | hCoV-19/USA/CA-ALSR-0812-SAN/2020\|EPI_ISL_483375\|2 | .................... | .................... | .................... | .................... | .................... | .................... | |
| E | 6743 | hCoV-19/USA/CA-ALSR-0813-SAN/2020\|EPI_ISL_483376\|2 | .................... | .................... | .................... | .................... | .................... | .................... | |
| E | 6744 | hCoV-19/USA/CA-ALSR-0815-SAN/2020\|EPI_ISL_483377\|2 | .................... | .................... | .................... | .................... | .................... | .................... | |
| E | 6745 | hCoV-19/USA/CA-ALSR-0816-SAN/2020\|EPI_ISL_483378\|2 | .................... | .................... | .................... | .................... | .................... | .................... | |
| E | 6746 | hCoV-19/USA/CA-ALSR-0820-SAN/2020\|EPI_ISL_483380\|2 | .................... | .................... | .................... | .................... | .................... | .................... | |
| E | 6747 | hCoV-19/USA/CA-ALSR-0821-SAN/2020\|EPI_ISL_483381\|2 | .................... | .................... | .................... | .................... | .................... | .................... | |
| E | 6748 | hCoV-19/USA/CA-ALSR-0822-SAN/2020\|EPI_ISL_483382\|2 | .................... | .................... | .................... | .................... | .................... | .................... | |
| E | 6749 | hCoV-19/USA/CA-ALSR-0823-SAN/2020\|EPI_ISL_483383\|2 | .................... | .................... | .................... | .................... | .................... | .................... | |
| E | 6750 | hCoV-19/USA/CA-ALSR-0827-SAN/2020\|EPI_ISL_483384\|2 | .................... | .................... | .................... | .................... | .................... | .................... | |
| E | 6751 | hCoV-19/USA/CA-ALSR-0828-IPL/2020\|EPI_ISL_483385\|2 | .................... | .................... | .................... | .................... | .................... | .................... | |
| E | 6752 | hCoV-19/USA/CA-ALSR-0830-SAN/2020\|EPI_ISL_483386\|2 | .................... | .................... | .................... | .................... | .................... | .................... | |
| E | 6753 | hCoV-19/USA/CA-ALSR-0831-IPL/2020\|EPI_ISL_483387\|2 | .................... | .................... | .................... | .................... | .................... | .................... | |
| E | 6754 | hCoV-19/USA/CA-ALSR-0832-SAN/2020\|EPI_ISL_483388\|2 | .................... | .................... | .................... | .................... | .................... | .................... | |
| E | 6755 | hCoV-19/USA/CA-ALSR-0833-SAN/2020\|EPI_ISL_483389\|2 | .................... | .................... | .................... | .................... | .................... | .................... | |
| E | 6756 | hCoV-19/USA/CA-ALSR-0834-IPL/2020\|EPI_ISL_483390\|2 | .................... | .................... | .................... | .................... | .................... | .................... | |
| E | 6757 | hCoV-19/USA/CA-ALSR-0837-SAN/2020\|EPI_ISL_483392\|2 | .................... | .................... | .................... | .................... | .................... | .................... | |
| E | 6758 | hCoV-19/USA/CA-ALSR-0839-IPL/2020\|EPI_ISL_483393\|2 | .................... | .................... | .................... | .................... | .................... | .................... | |
| E | 6759 | hCoV-19/USA/CA-ALSR-0843-IPL/2020\|EPI_ISL_483394\|2 | .................... | .A.................. | .................... | .................... | .................... | .................... | |
| E | 6760 | hCoV-19/USA/CA-ALSR-0845-IPL/2020\|EPI_ISL_483395\|2 | .................... | .A.................. | .................... | .................... | .................... | .................... | |
| E | 6761 | hCoV-19/USA/CA-ALSR-0847-IPL/2020\|EPI_ISL_483396\|2 | .................... | .................... | .................... | .................... | .................... | .................... | |
| E | 6762 | hCoV-19/USA/CA-ALSR-0849-SAN/2020\|EPI_ISL_483397\|2 | .................... | .................... | .................... | .................... | .................... | .................... | |
| E | 6763 | hCoV-19/USA/CA-ALSR-0850-NBG/2020\|EPI_ISL_483398\|2 | .................... | .................... | .................... | .................... | .................... | .................... | |
| E | 6764 | hCoV-19/USA/CA-ALSR-0852-NBG/2020\|EPI_ISL_483399\|2 | .................... | .................... | .................... | .................... | .................... | .................... | |
| E | 6765 | hCoV-19/Scotland/CVR2711/2020\|EPI_ISL_484651\|2020- | .................... | ...................A | .................... | .................... | .................... | .................... | 1 |
| E | 6766 | hCoV-19/Scotland/CVR2908/2020\|EPI_ISL_484652\|2020- | .................... | .A.................. | .................... | .................... | .................... | .................... | |
| E | 6767 | hCoV-19/Scotland/CVR2910/2020\|EPI_ISL_484653\|2020- | .................... | .................... | .................... | .................... | .................... | .................... | |
| E | 6768 | hCoV-19/Scotland/CVR3001/2020\|EPI_ISL_484654\|2020- | .................... | .................... | .................... | .................... | .................... | .................... | |
| E | 6769 | hCoV-19/Scotland/CVR3003/2020\|EPI_ISL_484655\|2020- | .................... | .A.................. | .................... | .................... | .................... | .................... | |
| E | 6770 | hCoV-19/Scotland/CVR3007/2020\|EPI_ISL_484656\|2020- | .................... | .................... | .................... | .................... | .................... | .................... | |
| E | 6771 | hCoV-19/Scotland/CVR3504/2020\|EPI_ISL_484657\|2020- | .................... | .................... | .................... | .................... | .................... | .................... | |
| E | 6772 | hCoV-19/Scotland/CVR3851/2020\|EPI_ISL_484658\|2020- | .................... | .................... | .................... | .................... | .................... | .................... | |
| E | 6773 | hCoV-19/Scotland/CVR3855/2020\|EPI_ISL_484659\|2020- | .................... | .................... | .................... | .................... | .................... | .................... | |
| E | 6774 | hCoV-19/Scotland/CVR3857/2020\|EPI_ISL_484660\|2020- | .................... | .A.................. | .................... | .................... | .................... | .................... | |
| E | 6775 | hCoV-19/Scotland/CVR3861/2020\|EPI_ISL_484661\|2020- | .................... | .................... | .................... | .................... | .................... | .................... | |
| E | 6776 | hCoV-19/Scotland/CVR3868/2020\|EPI_ISL_484662\|2020- | .................... | ...................A | .................... | .................... | .................... | .................... | 1 |
| E | 6777 | hCoV-19/Scotland/CVR3869/2020\|EPI_ISL_484663\|2020- | .................... | ...................A | .................... | .................... | .................... | .................... | 1 |
| E | 6778 | hCoV-19/Scotland/CVR3870/2020\|EPI_ISL_484664\|2020- | .................... | ...................A | .................... | .................... | .................... | .................... | 1 |
| E | 6779 | hCoV-19/Scotland/CVR3876/2020\|EPI_ISL_484665\|2020- | .................... | ...................A | .................... | .................... | .................... | .................... | 1 |
| E | 6780 | hCoV-19/Scotland/CVR3877/2020\|EPI_ISL_484666\|2020- | .................... | .................... | .................... | .................... | .................... | .................... | |
| E | 6781 | hCoV-19/Scotland/CVR3878/2020\|EPI_ISL_484667\|2020- | .................... | .................... | .................... | .................... | .................... | .................... | |
| E | 6782 | hCoV-19/Scotland/CVR3880/2020\|EPI_ISL_484668\|2020- | .................... | ...................A | .................... | .................... | .................... | .................... | 1 |
| E | 6783 | hCoV-19/Scotland/CVR3882/2020\|EPI_ISL_484669\|2020- | .................... | .................... | .................... | .................... | .................... | .................... | |
| E | 6784 | hCoV-19/Scotland/CVR3883/2020\|EPI_ISL_484670\|2020- | .................... | .................... | .................... | .................... | .................... | .................... | |
| E | 6785 | hCoV-19/Scotland/CVR3884/2020\|EPI_ISL_484671\|2020- | .................... | .................... | .................... | .................... | .................... | .................... | |
| E | 6786 | hCoV-19/Scotland/CVR3886/2020\|EPI_ISL_484672\|2020- | .................... | .................... | .................... | .................... | .................... | .................... | |
| E | 6787 | hCoV-19/Scotland/CVR3888/2020\|EPI_ISL_484673\|2020- | .................... | .................... | .................... | .................... | .................... | .................... | |
| E | 6788 | hCoV-19/Scotland/CVR3889/2020\|EPI_ISL_484674\|2020- | .................... | ...................A | .................... | .................... | .................... | .................... | 1 |
| E | 6789 | hCoV-19/Scotland/CVR3890/2020\|EPI_ISL_484675\|2020- | .................... | .................... | .................... | .................... | .................... | .................... | |
| E | 6790 | hCoV-19/Scotland/CVR3892/2020\|EPI_ISL_484676\|2020- | .................... | .................... | .................... | .................... | .................... | .................... | |
| E | 6791 | hCoV-19/Scotland/CVR3895/2020\|EPI_ISL_484677\|2020- | .................... | .................... | .................... | .................... | .................... | .................... | |
| E | 6792 | hCoV-19/Scotland/CVR3896/2020\|EPI_ISL_484678\|2020- | .................... | .................... | .................... | .................... | .................... | .................... | |
| E | 6793 | hCoV-19/Scotland/CVR3897/2020\|EPI_ISL_484679\|2020- | .................... | .................... | .................... | .................... | .................... | .................... | |
| E | 6794 | hCoV-19/Scotland/EDB6603/2020\|EPI_ISL_484680\|2020- | .................... | .................... | .................... | .................... | .................... | .................... | |
| E | 6795 | hCoV-19/Scotland/EDB6604/2020\|EPI_ISL_484681\|2020- | .................... | .................... | .................... | .................... | .................... | .................... | |
| E | 6796 | hCoV-19/Scotland/EDB6607/2020\|EPI_ISL_484682\|2020- | .................... | .................... | .................... | .................... | .................... | .................... | |
| E | 6797 | hCoV-19/Scotland/EDB6608/2020\|EPI_ISL_484683\|2020- | .................... | .................... | .................... | .................... | .................... | .................... | |
| E | 6798 | hCoV-19/Belgium/ULG-10208/2020\|EPI_ISL_484697\|2020 | .................... | .................... | .................... | .................... | .................... | .................... | |
| E | 6799 | hCoV-19/Belgium/ULG-10209/2020\|EPI_ISL_484698\|2020 | .................... | .................... | .................... | .................... | .................... | .................... | |
| E | 6800 | hCoV-19/Belgium/ULG-10210/2020\|EPI_ISL_484699\|2020 | .................... | .................... | .................... | .................... | .................... | .................... | |
| E | 6801 | hCoV-19/USA/WA-UW-2305/2020\|EPI_ISL_485982\|2020-03 | .................... | .................... | .................... | .................... | .................... | .................... | |
| E | 6802 | hCoV-19/USA/WA-UW-2344/2020\|EPI_ISL_485983\|2020-04 | .................... | .................... | .................... | .................... | .................... | .................... | |
| E | 6803 | hCoV-19/USA/WA-UW-2346/2020\|EPI_ISL_485984\|2020-04 | .................... | .................... | .................... | .................... | .................... | .................... | |
| E | 6804 | hCoV-19/USA/WA-UW-2361/2020\|EPI_ISL_485985\|2020-04 | .................... | .................... | .................... | .................... | .................... | .................... | |
| E | 6805 | hCoV-19/USA/WA-UW-2368/2020\|EPI_ISL_485986\|2020-03 | .................... | .................... | .................... | .................... | .................... | .................... | |
| E | 6806 | hCoV-19/USA/WA-UW-2369/2020\|EPI_ISL_485987\|2020-03 | .................... | .................... | .................... | .................... | .................... | .................... | |
| E | 6807 | hCoV-19/USA/WA-UW-2377/2020\|EPI_ISL_485988\|2020-03 | .................... | .................... | .................... | .................... | .................... | .................... | |
| E | 6808 | hCoV-19/USA/WA-UW-2382/2020\|EPI_ISL_485989\|2020-03 | .................... | .................... | .................... | .................... | .................... | .................... | |
| E | 6809 | hCoV-19/USA/WA-UW-2414/2020\|EPI_ISL_485990\|2020-04 | .................... | .................... | .................... | .................... | .................... | .................... | |
| E | 6810 | hCoV-19/USA/WA-UW-2428/2020\|EPI_ISL_485992\|2020-04 | .................... | .................... | .................... | .................... | .................... | .................... | |
| E | 6811 | hCoV-19/USA/WA-UW-2430/2020\|EPI_ISL_485993\|2020-04 | .................... | .................... | .................... | .................... | .................... | .................... | |
| E | 6812 | hCoV-19/USA/WA-UW-2526/2020\|EPI_ISL_485994\|2020-03 | .................... | .................... | .................... | .................... | .................... | .................... | |
| E | 6813 | hCoV-19/USA/WA-UW-2532/2020\|EPI_ISL_485995\|2020-03 | .................... | .................... | .................... | .................... | .................... | .................... | |
| E | 6814 | hCoV-19/USA/WA-UW-2537/2020\|EPI_ISL_485996\|2020-03 | .................... | .................... | .................... | .................... | .................... | .................... | |
| E | 6815 | hCoV-19/USA/WA-UW-2540/2020\|EPI_ISL_485997\|2020-03 | .................... | .................... | .................... | .................... | .................... | .................... | |
| E | 6816 | hCoV-19/USA/WA-UW-2551/2020\|EPI_ISL_485998\|2020-03 | .................... | .................... | .................... | .................... | .................... | .................... | |
| E | 6817 | hCoV-19/USA/WA-UW-2558/2020\|EPI_ISL_485999\|2020-03 | .................... | .................... | .................... | .................... | .................... | .................... | |

| | | | | | | | | | |
|---|---|---|---|---|---|---|---|---|---|
| E | 6818 hCoV-19/England/CAMB-1ACB13/2020\|EPI_ISL_489500\|20 | ···················· | ···················· | ···················· | ···················· | ···················· | ···················· | | |
| E | 6819 hCoV-19/Wales/PHWC-166474/2020\|EPI_ISL_494001\|2020 | ···················· | ···················· | ···················· | ···················· | ···················· | ···················· | | |
| E | 6821 hCoV-19/Wales/PHWC-1664A1/2020\|EPI_ISL_494003\|2020 | ···················· | ···················· | ···················· | ···················· | ···················· | ···················· | | |
| E | 6822 hCoV-19/Wales/PHWC-1664B0/2020\|EPI_ISL_494004\|2020 | ···················· | ···················· | ···················· | ···················· | ···················· | ···················· | | |
| E | 6823 hCoV-19/Wales/PHWC-1664CF/2020\|EPI_ISL_494005\|2020 | ···················· | ···················· | ···················· | ···················· | ···················· | ···················· | | |
| E | 6824 hCoV-19/Wales/PHWC-1664DE/2020\|EPI_ISL_494006\|2020 | ···················· | ···················· | ···················· | ···················· | ···················· | ···················· | | |
| E | 6825 hCoV-19/Wales/PHWC-1664FC/2020\|EPI_ISL_494008\|2020 | ···················· | ···················· | ···················· | ···················· | ···················· | ···················· | | |
| E | 6826 hCoV-19/Wales/PHWC-166508/2020\|EPI_ISL_494009\|2020 | ···················· | ···················· | ···················· | ···················· | ···················· | ···················· | | |
| E | 6827 hCoV-19/Wales/PHWC-166517/2020\|EPI_ISL_494010\|2020 | ···················· | ···················· | ···················· | ···················· | ···················· | ···················· | | |
| E | 6832 hCoV-19/Wales/PHWC-166562/2020\|EPI_ISL_494015\|2020 | ···················· | ···················· | ···················· | ···················· | ···················· | ···················· | | |
| E | 6833 hCoV-19/Wales/PHWC-166580/2020\|EPI_ISL_494016\|2020 | ···················· | ···················· | ···················· | ···················· | ···················· | ···················· | | |
| E | 6836 hCoV-19/Wales/PHWC-1665DB/2020\|EPI_ISL_494021\|2020 | ···················· | ···················· | ···················· | ···················· | ···················· | ···················· | | |
| E | 6837 hCoV-19/Wales/PHWC-166632/2020\|EPI_ISL_494024\|2020 | ···················· | ···················· | ···················· | ···················· | ···················· | ···················· | | |
| E | 6838 hCoV-19/Wales/PHWC-166650/2020\|EPI_ISL_494025\|2020 | ···················· | ···················· | ···················· | ···················· | ···················· | ···················· | | |
| E | 6839 hCoV-19/Wales/PHWC-16666F/2020\|EPI_ISL_494026\|2020 | ···················· | ···················· | ···················· | ···················· | ···················· | ···················· | | |
| E | 6840 hCoV-19/Wales/PHWC-16667E/2020\|EPI_ISL_494027\|2020 | ···················· | ···················· | ···················· | ···················· | ···················· | ···················· | | |
| E | 6841 hCoV-19/Wales/PHWC-16668D/2020\|EPI_ISL_494028\|2020 | ···················· | ···················· | ···················· | ···················· | ···················· | ···················· | | |
| E | 6842 hCoV-19/Wales/PHWC-1666BA/2020\|EPI_ISL_494030\|2020 | ···················· | ···················· | ···················· | ···················· | ···················· | ·NNN················ | | 1 |
| E | 6843 hCoV-19/Wales/PHWC-166702/2020\|EPI_ISL_494031\|2020 | ···················· | ···················· | ···················· | ···················· | ···················· | ···················· | | |
| E | 6846 hCoV-19/Wales/PHWC-16675D/2020\|EPI_ISL_494035\|2020 | ···················· | ···················· | ···················· | ···················· | ···················· | ···················· | | |
| E | 6847 hCoV-19/Wales/PHWC-16676C/2020\|EPI_ISL_494036\|2020 | ···················· | ···················· | ···················· | ···················· | ·NN················· | ···················· | | |
| E | 6849 hCoV-19/Wales/PHWC-1667C6/2020\|EPI_ISL_494040\|2020 | ···················· | ···················A | ···················· | ···················· | ···················· | ···················· | 1 | |
| E | 6850 hCoV-19/Wales/PHWC-1667D5/2020\|EPI_ISL_494041\|2020 | ···················· | ···················· | ···················· | ···················· | ···················· | ···················· | | |
| E | 6851 hCoV-19/Wales/PHWC-1667F3/2020\|EPI_ISL_494043\|2020 | ···················· | ···················· | ···················· | ···················· | ···················· | ···················· | | |
| E | 6852 hCoV-19/Wales/PHWC-16680F/2020\|EPI_ISL_494044\|2020 | ···················· | ···················· | ···················· | ···················· | ···················· | ···················· | | |
| E | 6853 hCoV-19/Wales/PHWC-16681E/2020\|EPI_ISL_494045\|2020 | ···················· | ···················· | ···················· | ···················· | NNN················· | ···················· | | 1 |
| E | 6855 hCoV-19/Wales/PHWC-16683C/2020\|EPI_ISL_494047\|2020 | ···················· | ···················· | ···················· | ···················· | ···················· | ···················· | | |
| E | 6856 hCoV-19/Wales/PHWC-16685A/2020\|EPI_ISL_494049\|2020 | ···················· | ···················· | ···················· | ···················· | ···················· | ···················· | | |
| E | 6857 hCoV-19/Wales/PHWC-166869/2020\|EPI_ISL_494050\|2020 | ···················· | ···················· | ···················· | ···················· | ···················· | ···················· | | |
| E | 6858 hCoV-19/Wales/PHWC-166878/2020\|EPI_ISL_494051\|2020 | ···················· | ···A················ | ···················· | ···················· | ···················· | ···················· | | |
| E | 6859 hCoV-19/Wales/PHWC-166887/2020\|EPI_ISL_494052\|2020 | ···················· | ···················· | ···················· | ···················· | ·NNN················ | ···················· | | 1 |
| E | 6860 hCoV-19/Wales/PHWC-166896/2020\|EPI_ISL_494053\|2020 | ···················· | ···················· | ···················· | ···················· | ···················· | ···················· | | |
| E | 6861 hCoV-19/Wales/PHWC-1668C3/2020\|EPI_ISL_494055\|2020 | ···················· | ···················· | ···················· | ···················· | ···················· | ···················· | | |
| E | 6862 hCoV-19/Wales/PHWC-166ACD/2020\|EPI_ISL_494078\|2020 | ···················· | ···················· | ···················· | ···················· | ···················· | ···················· | | |
| E | 6863 hCoV-19/Wales/PHWC-166ADC/2020\|EPI_ISL_494079\|2020 | ···················· | ···················· | ···················· | ···················· | ···················· | ···················· | | |
| E | 6864 hCoV-19/Wales/PHWC-166AEB/2020\|EPI_ISL_494080\|2020 | ···················· | ···················· | ···················· | ···················· | ···················· | ···················· | | |
| E | 6865 hCoV-19/Wales/PHWC-166B06/2020\|EPI_ISL_494082\|2020 | ···················· | ···················· | ···················· | ···················· | ···················· | ···················· | | |
| E | 6866 hCoV-19/Wales/PHWC-166B33/2020\|EPI_ISL_494084\|2020 | ···················· | ···················· | ···················· | ···················· | ···············NN··· | ···················· | | |
| E | 6867 hCoV-19/Wales/PHWC-166B42/2020\|EPI_ISL_494085\|2020 | ···················· | ···················· | ···················· | ···················· | ···················· | ···················· | | |
| E | 6869 hCoV-19/Wales/PHWC-166B7F/2020\|EPI_ISL_494087\|2020 | ···················· | ···················· | ···················· | ···················· | ···················· | ···················· | | |
| E | 6870 hCoV-19/Wales/PHWC-166B8E/2020\|EPI_ISL_494088\|2020 | ···················· | ···················· | ···················· | ···················· | ···················· | ···················· | | |
| E | 6871 hCoV-19/USA/VA-DCLS-0627/2020\|EPI_ISL_495300\|2020- | ···················· | ···················· | ···················· | ···················· | ···················· | ···················· | | |
| E | 6872 hCoV-19/USA/VA-DCLS-0628/2020\|EPI_ISL_495301\|2020- | ···················· | ···················· | ···················· | ···················· | ···················· | ···················· | | |
| E | 6873 hCoV-19/USA/VA-DCLS-0629/2020\|EPI_ISL_495302\|2020- | ···················· | ···················· | ···················· | ···················· | ···················· | ···················· | | |
| E | 6874 hCoV-19/USA/VA-DCLS-0630/2020\|EPI_ISL_495303\|2020- | ···················· | ···················· | ···················· | ···················· | ···················· | ···················· | | |
| E | 6875 hCoV-19/USA/VA-DCLS-0631/2020\|EPI_ISL_495304\|2020- | ···················· | ···················· | ···················· | ···················· | ···················· | ···················· | | |
| E | 6876 hCoV-19/USA/VA-DCLS-0632/2020\|EPI_ISL_495305\|2020- | ···················· | ···················· | ···················· | ···················· | ···················· | ···················· | | |
| E | 6877 hCoV-19/USA/VA-DCLS-0633/2020\|EPI_ISL_495306\|2020- | ···················· | ···················· | ···················· | ···················· | ···················· | ···················· | | |
| E | 6878 hCoV-19/USA/VA-DCLS-0635/2020\|EPI_ISL_495307\|2020- | ···················· | ···················· | ···················· | ···················· | ···················· | ···················· | | |
| E | 6879 hCoV-19/USA/VA-DCLS-0636/2020\|EPI_ISL_495308\|2020- | ···················· | ···················· | ···················· | ···················· | ···················· | ···················· | | |
| E | 6880 hCoV-19/USA/VA-DCLS-0637/2020\|EPI_ISL_495309\|2020- | ···················· | ···················· | ···················· | ···················· | ···················· | ···················· | | |
| E | 6881 hCoV-19/USA/VA-DCLS-0638/2020\|EPI_ISL_495310\|2020- | ···················· | ···················· | ···················· | ···················· | ···················· | ···················· | | |
| E | 6882 hCoV-19/USA/VA-DCLS-0639/2020\|EPI_ISL_495311\|2020- | ···················· | ···················· | ···················· | ···················· | ···················· | ···················· | | |
| E | 6883 hCoV-19/USA/VA-DCLS-0640/2020\|EPI_ISL_495312\|2020- | ···················· | ···················· | ···················· | ···················· | ···················· | ···················· | | |
| E | 6884 hCoV-19/USA/VA-DCLS-0641/2020\|EPI_ISL_495313\|2020- | ···················· | ···················· | ···················· | ···················· | ···················· | ···················· | | |
| E | 6885 hCoV-19/USA/VA-DCLS-0642/2020\|EPI_ISL_495314\|2020- | ···················· | ···················· | ···················· | ···················· | ···················· | ···················· | | |
| E | 6886 hCoV-19/USA/VA-DCLS-0643/2020\|EPI_ISL_495315\|2020- | ···················· | ···················· | ···················· | ···················· | ···················· | ···················· | | |
| E | 6887 hCoV-19/USA/VA-DCLS-0644/2020\|EPI_ISL_495316\|2020- | ···················· | ···················· | ···················· | ···················· | ···················· | ···················· | | |
| E | 6888 hCoV-19/USA/VA-DCLS-0645/2020\|EPI_ISL_495317\|2020- | ···················· | ···················· | ···················· | ···················· | ···················· | ···················· | | |
| E | 6889 hCoV-19/USA/VA-DCLS-0647/2020\|EPI_ISL_495319\|2020- | ···················· | ···················· | ···················· | ···················· | ···················· | ···················· | | |
| E | 6890 hCoV-19/USA/VA-DCLS-0648/2020\|EPI_ISL_495320\|2020- | ···················· | ···················· | ···················· | ···················· | ···················· | ···················· | | |
| E | 6891 hCoV-19/USA/VA-DCLS-0651/2020\|EPI_ISL_495323\|2020- | ···················· | ···················· | ···················· | ···················· | ···················· | ···················· | | |
| E | 6892 hCoV-19/USA/VA-DCLS-0652/2020\|EPI_ISL_495324\|2020- | ···················· | ···················· | ···················· | ···················· | ···················· | ···················· | | |
| E | 6893 hCoV-19/USA/FL-BPHL-0426/2020\|EPI_ISL_495332\|2020- | ···················· | ···················· | ···················· | ···················· | ···················· | ···················· | | |
| E | 6894 hCoV-19/USA/FL-BPHL-0427/2020\|EPI_ISL_495333\|2020- | ···················· | ···················· | ···················· | ···················· | ···················· | ···················· | | |
| E | 6895 hCoV-19/USA/FL-BPHL-0428/2020\|EPI_ISL_495334\|2020- | ···················· | ···················· | ···················· | ···················· | ···················· | ···················· | | |
| E | 6896 hCoV-19/USA/FL-BPHL-0436/2020\|EPI_ISL_495342\|2020- | ···················· | ···················· | ···················· | ···················· | ···················· | ···················· | | |
| E | 6897 hCoV-19/USA/FL-BPHL-0438/2020\|EPI_ISL_495344\|2020- | ···················· | ···················· | ···················· | ···················· | ···················· | ···················· | | |
| E | 6899 hCoV-19/USA/FL-BPHL-0441/2020\|EPI_ISL_495347\|2020- | ···················· | ···················· | ···················· | ···················· | ···················· | ···················· | | |
| E | 6901 hCoV-19/USA/FL-BPHL-0444/2020\|EPI_ISL_495350\|2020- | ···················· | ···················· | ···················· | ···················· | ···················· | ···················· | | |
| E | 6902 hCoV-19/USA/FL-BPHL-0445/2020\|EPI_ISL_495351\|2020- | ···················· | ···················· | ···················· | ···················· | ···················· | ···················· | | |
| E | 6903 hCoV-19/USA/FL-BPHL-0449/2020\|EPI_ISL_495355\|2020- | ···················· | ···················· | ···················· | ···················· | ···················· | ···················· | | |
| E | 6904 hCoV-19/USA/FL-BPHL-0451/2020\|EPI_ISL_495357\|2020- | ···················· | ···················· | ···················· | ···················· | ···················· | ···················· | | |
| E | 6905 hCoV-19/USA/FL-BPHL-0453/2020\|EPI_ISL_495359\|2020- | ···················· | ···················· | ···················· | ···················· | ···················· | ···················· | | |
| E | 6906 hCoV-19/USA/FL-BPHL-0454/2020\|EPI_ISL_495360\|2020- | ···················· | ···················· | ···················· | ···················· | ···················· | ···················· | | |
| E | 6907 hCoV-19/USA/FL-BPHL-0457/2020\|EPI_ISL_495363\|2020- | ···················· | ···················· | ···················· | ···················· | ···················· | ···················· | | |
| E | 6908 hCoV-19/USA/FL-BPHL-0465/2020\|EPI_ISL_495371\|2020- | ···················· | ···················· | ···················· | ···················· | ···················· | ···················· | | |
| E | 6909 hCoV-19/USA/FL-BPHL-0472/2020\|EPI_ISL_495378\|2020- | ···················· | ···················· | ···················· | ···················· | ···················· | ···················· | | |
| E | 6910 hCoV-19/USA/FL-BPHL-0473/2020\|EPI_ISL_495379\|2020- | ···················· | ···················· | ···················· | ···················· | ···················· | ···················· | | |
| E | 6911 hCoV-19/USA/FL-BPHL-0478/2020\|EPI_ISL_495384\|2020- | ···················· | ···················· | ···················· | ···················· | ···················· | ···················· | | |

| | | | | | | | | |
|---|---|---|---|---|---|---|---|---|
| E | 6912 hCoV-19/USA/FL-BPHL-0480/2020\|EPI_ISL_495386\|2020- | .................... | .................... | .................... | .................... | .................... | .................... | |
| E | 6913 hCoV-19/England/BRIS-12E302/2020\|EPI_ISL_482025\|20 | .................... | .................... | .................... | .................... | .................... | .................... | |
| E | 6914 hCoV-19/England/BRIS-12DDC9/2020\|EPI_ISL_482026\|20 | .................... | .................... | .................... | .................... | .................... | .................... | |
| E | 6915 hCoV-19/England/BRIS-12E36C/2020\|EPI_ISL_482027\|20 | .................... | .................... | .................... | .................... | .................... | .................... | |
| E | 6916 hCoV-19/England/BRIS-12E41E/2020\|EPI_ISL_482028\|20 | .................... | .................... | .................... | .................... | .................... | .................... | |
| E | 6917 hCoV-19/England/BRIS-12E162/2020\|EPI_ISL_482030\|20 | .................... | .................... | .................... | .................... | .................... | .................... | |
| E | 6918 hCoV-19/USA/CA-ALSR-0684-SAN/2020\|EPI_ISL_483300\|2 | .................... | .................... | .................... | .................... | .................... | .................... | |
| E | 6919 hCoV-19/USA/CA-ALSR-0685-SAN/2020\|EPI_ISL_483301\|2 | .................... | .................... | .................... | .................... | .................... | .................... | |
| E | 6920 hCoV-19/USA/CA-ALSR-0686-SAN/2020\|EPI_ISL_483302\|2 | .................... | .................... | .................... | .................... | .................... | .................... | |
| E | 6921 hCoV-19/USA/CA-ALSR-0687-SAN/2020\|EPI_ISL_483303\|2 | .................... | .................... | .................... | .................... | .................... | .................... | |
| E | 6922 hCoV-19/USA/CA-ALSR-0689-SAN/2020\|EPI_ISL_483304\|2 | .................... | .................... | .................... | .................... | .................... | .................... | |
| E | 6923 hCoV-19/USA/CA-ALSR-0690-SAN/2020\|EPI_ISL_483305\|2 | .................... | .A.................. | .................... | .................... | .................... | .................... | |
| E | 6924 hCoV-19/USA/CA-ALSR-0696-SAN/2020\|EPI_ISL_483307\|2 | .................... | .................... | .................... | .................... | .................... | .................... | |
| E | 6925 hCoV-19/USA/CA-ALSR-0697-SAN/2020\|EPI_ISL_483308\|2 | .................... | .................... | .................... | .................... | .................... | .................... | |
| E | 6926 hCoV-19/USA/CA-ALSR-0698-SAN/2020\|EPI_ISL_483309\|2 | .................... | .................... | .................... | .................... | .................... | .................... | |
| E | 6927 hCoV-19/USA/CA-ALSR-0699-SAN/2020\|EPI_ISL_483310\|2 | .................... | .................... | .................... | .................... | .................... | .................... | |
| E | 6928 hCoV-19/USA/CA-ALSR-0700-SAN/2020\|EPI_ISL_483311\|2 | .................... | .................... | .................... | .................... | .................... | .................... | |
| E | 6929 hCoV-19/USA/CA-ALSR-0701-SAN/2020\|EPI_ISL_483312\|2 | .................... | .................... | .................... | .................... | .................... | .................... | |
| E | 6930 hCoV-19/USA/CA-ALSR-0704-SAN/2020\|EPI_ISL_483313\|2 | .................... | .................... | .................... | .................... | .................... | .................... | |
| E | 6931 hCoV-19/USA/CA-ALSR-0706-SAN/2020\|EPI_ISL_483315\|2 | .................... | .................... | .................... | .................... | .................... | .................... | |
| E | 6932 hCoV-19/USA/CA-ALSR-0707-SAN/2020\|EPI_ISL_483316\|2 | .................... | .................... | .................... | .................... | .................... | .................... | |
| E | 6933 hCoV-19/USA/CA-ALSR-0709-SAN/2020\|EPI_ISL_483317\|2 | .................... | .................... | .................... | .................... | .................... | .................... | |
| E | 6934 hCoV-19/USA/CA-ALSR-0711-SAN/2020\|EPI_ISL_483318\|2 | .................... | .................... | .................... | .................... | .................... | .................... | |
| E | 6935 hCoV-19/USA/CA-ALSR-0713-SAN/2020\|EPI_ISL_483319\|2 | .................... | .................... | .................... | .................... | .................... | .................... | |
| E | 6936 hCoV-19/USA/CA-ALSR-0781-SAN/2020\|EPI_ISL_483353\|2 | .................... | .................... | .................... | .................... | .................... | .................... | |
| E | 6937 hCoV-19/USA/CA-ALSR-0782-SAN/2020\|EPI_ISL_483354\|2 | .................... | .................... | .................... | .................... | .................... | .................... | |
| E | 6938 hCoV-19/USA/CA-ALSR-0783-SAN/2020\|EPI_ISL_483355\|2 | .................... | .................... | .................... | .................... | .................... | .................... | |
| E | 6939 hCoV-19/USA/CA-ALSR-0786-IPL/2020\|EPI_ISL_483356\|2 | .................... | .................... | .................... | .................... | .................... | .................... | |
| E | 6940 hCoV-19/USA/CA-ALSR-0787-SAN/2020\|EPI_ISL_483357\|2 | .................... | .................... | .................... | .................... | .................... | .................... | |
| E | 6941 hCoV-19/USA/CA-ALSR-0788-SAN/2020\|EPI_ISL_483358\|2 | .................... | .................... | .................... | .................... | .................... | .................... | |
| E | 6942 hCoV-19/USA/CA-ALSR-0789-SAN/2020\|EPI_ISL_483359\|2 | .................... | .................... | .................... | .................... | .................... | .................... | |
| E | 6943 hCoV-19/USA/CA-ALSR-0790-SAN/2020\|EPI_ISL_483360\|2 | .................... | .................... | .................... | .................... | .................... | .................... | |
| E | 6944 hCoV-19/USA/CA-ALSR-0794-SAN/2020\|EPI_ISL_483362\|2 | .................... | .................... | .................... | .................... | .................... | .................... | |
| E | 6945 hCoV-19/USA/CA-ALSR-0795-SAN/2020\|EPI_ISL_483363\|2 | .................... | .................... | .................... | .................... | .................... | .................... | |
| E | 6946 hCoV-19/Scotland/CVR2551/2020\|EPI_ISL_484600\|2020- | .................... | .................... | .................... | .................... | .................... | .................... | |
| E | 6947 hCoV-19/Scotland/CVR2552/2020\|EPI_ISL_484601\|2020- | .................... | .................... | ..........T.T....... | .................... | .................... | .................... | |
| E | 6948 hCoV-19/Scotland/CVR2567/2020\|EPI_ISL_484603\|2020- | .................... | .A.................. | .................... | .................... | .................... | .................... | |
| E | 6949 hCoV-19/Scotland/CVR2569/2020\|EPI_ISL_484604\|2020- | .................... | .A.................. | .................... | .................... | .................... | .................... | |
| E | 6950 hCoV-19/Scotland/CVR2570/2020\|EPI_ISL_484605\|2020- | .................... | ...................A | .................... | .................... | .................... | .................... | 1 |
| E | 6951 hCoV-19/Scotland/CVR2581/2020\|EPI_ISL_484607\|2020- | .................... | .................... | .................... | .................... | .................... | .................... | |
| E | 6952 hCoV-19/Scotland/CVR2585/2020\|EPI_ISL_484608\|2020- | .................... | ...................A | .................... | .................... | .................... | .................... | 1 |
| E | 6953 hCoV-19/Scotland/CVR2589/2020\|EPI_ISL_484609\|2020- | .................... | .................... | .................... | .................... | .................... | .................... | |
| E | 6954 hCoV-19/Scotland/CVR2590/2020\|EPI_ISL_484610\|2020- | .................... | .................... | .................... | .................... | .................... | .................... | |
| E | 6955 hCoV-19/Scotland/CVR2598/2020\|EPI_ISL_484611\|2020- | .................... | .................... | .................... | .................... | .................... | .................... | |
| E | 6956 hCoV-19/Scotland/CVR2608/2020\|EPI_ISL_484612\|2020- | .................... | .A.................. | .................... | .................... | .................... | .................... | |
| E | 6957 hCoV-19/Scotland/CVR2612/2020\|EPI_ISL_484614\|2020- | .................... | .................... | .................... | .................... | .................... | .................... | |
| E | 6958 hCoV-19/Scotland/CVR2614/2020\|EPI_ISL_484615\|2020- | .................... | .................... | .................... | .................... | .................... | .................... | |
| E | 6959 hCoV-19/Scotland/CVR2615/2020\|EPI_ISL_484616\|2020- | .................... | .................... | .................... | .................... | .................... | .................... | |
| E | 6960 hCoV-19/Scotland/CVR2617/2020\|EPI_ISL_484617\|2020- | .................... | .................... | .................... | .................... | .................... | .................... | |
| E | 6961 hCoV-19/Scotland/CVR2621/2020\|EPI_ISL_484618\|2020- | .................... | .................... | .................... | .................... | .................... | .................... | |
| E | 6962 hCoV-19/Scotland/CVR2622/2020\|EPI_ISL_484619\|2020- | .................... | .................... | .................... | .................... | .................... | .................... | |
| E | 6963 hCoV-19/Scotland/CVR2623/2020\|EPI_ISL_484620\|2020- | .................... | .................... | .................... | .................... | .................... | .................... | |
| E | 6964 hCoV-19/Scotland/CVR2625/2020\|EPI_ISL_484621\|2020- | .................... | .................... | .................... | .................... | .................... | .................... | |
| E | 6965 hCoV-19/Scotland/CVR2630/2020\|EPI_ISL_484622\|2020- | .................... | ...................A | .................... | .................... | .................... | .................... | 1 |
| E | 6966 hCoV-19/Scotland/CVR2634/2020\|EPI_ISL_484623\|2020- | .................... | .................... | .................... | .................... | .................... | .................... | |
| E | 6967 hCoV-19/Scotland/CVR2636/2020\|EPI_ISL_484624\|2020- | .................... | .................... | .................... | .................... | .................... | .................... | |
| E | 6968 hCoV-19/Scotland/CVR2639/2020\|EPI_ISL_484625\|2020- | .................... | .................... | .................... | .................... | .................... | .................... | |
| E | 6969 hCoV-19/Scotland/CVR2642/2020\|EPI_ISL_484626\|2020- | .................... | ...................A | .................... | .................... | .................... | .................... | 1 |
| E | 6970 hCoV-19/Scotland/CVR2643/2020\|EPI_ISL_484627\|2020- | .................... | .................... | .................... | .................... | .................... | .................... | |
| E | 6971 hCoV-19/Scotland/CVR2648/2020\|EPI_ISL_484629\|2020- | .................... | .................... | .................... | .................... | .................... | .................... | |
| E | 6972 hCoV-19/Scotland/CVR2655/2020\|EPI_ISL_484630\|2020- | .................... | .................... | .................... | .................... | .................... | .................... | |
| E | 6973 hCoV-19/Scotland/CVR2665/2020\|EPI_ISL_484631\|2020- | .................... | .................... | .................... | .................... | .................... | .................... | |
| E | 6974 hCoV-19/Scotland/CVR2666/2020\|EPI_ISL_484632\|2020- | .................... | .A.................. | .................... | .................... | .................... | .................... | |
| E | 6975 hCoV-19/Scotland/CVR2671/2020\|EPI_ISL_484633\|2020- | .................... | .................... | .................... | .................... | .................... | .................... | |
| E | 6976 hCoV-19/Scotland/CVR2674/2020\|EPI_ISL_484634\|2020- | .................... | .A.................. | .................... | .................... | .................... | .................... | |
| E | 6977 hCoV-19/Scotland/CVR2677/2020\|EPI_ISL_484635\|2020- | .................... | .................... | .................... | .................... | .................... | .................... | |
| E | 6978 hCoV-19/Scotland/CVR2678/2020\|EPI_ISL_484636\|2020- | .................... | .................... | .................... | .................... | .................... | .................... | |
| E | 6979 hCoV-19/Scotland/CVR2683/2020\|EPI_ISL_484637\|2020- | .................... | .................... | .................... | .................... | .................... | .................... | |
| E | 6980 hCoV-19/Scotland/CVR2684/2020\|EPI_ISL_484638\|2020- | .................... | .................... | .................... | .................... | .................... | .................... | |
| E | 6981 hCoV-19/Scotland/CVR2685/2020\|EPI_ISL_484639\|2020- | .................... | ...................A | .................... | .................... | .................... | .................... | 1 |
| E | 6982 hCoV-19/Scotland/CVR2689/2020\|EPI_ISL_484640\|2020- | .................... | .................... | .................... | .................... | .................... | .................... | |
| E | 6983 hCoV-19/Scotland/CVR2693/2020\|EPI_ISL_484641\|2020- | .................... | .................... | .................... | .................... | .................... | .................... | |
| E | 6984 hCoV-19/Scotland/CVR2694/2020\|EPI_ISL_484642\|2020- | .................... | .................... | .................... | .................... | .................... | .................... | |
| E | 6985 hCoV-19/Scotland/CVR2695/2020\|EPI_ISL_484643\|2020- | .................... | .................... | .................... | .................... | .................... | .................... | |
| E | 6986 hCoV-19/Scotland/CVR2696/2020\|EPI_ISL_484644\|2020- | .................... | .................... | .................... | .................... | .................... | .................... | |
| E | 6987 hCoV-19/Scotland/CVR2697/2020\|EPI_ISL_484645\|2020- | .................... | .................... | .................... | .................... | .................... | .................... | |
| E | 6988 hCoV-19/Scotland/CVR2702/2020\|EPI_ISL_484646\|2020- | .................... | .................... | .................... | .................... | .................... | .................... | |
| E | 6989 hCoV-19/Scotland/CVR2705/2020\|EPI_ISL_484647\|2020- | .................... | .................... | .................... | .................... | .................... | .................... | |
| E | 6990 hCoV-19/Scotland/CVR2706/2020\|EPI_ISL_484648\|2020- | .................... | .................... | .................... | .................... | .................... | .................... | |
| E | 6991 hCoV-19/Scotland/CVR2707/2020\|EPI_ISL_484649\|2020- | .................... | .................... | .................... | .................... | .................... | .................... | |

| | | | | | | | | | |
|---|---|---|---|---|---|---|---|---|---|
| E | 6992 | hCoV-19/Scotland/CVR2709/2020\|EPI_ISL_484650\|2020- | ···················· | ···················A | ···················· | ···················· | ···················· | ···················· | 1 |
| E | 6993 | hCoV-19/Scotland/EDB6609/2020\|EPI_ISL_484684\|2020- | ···················· | ···················· | ···················· | ···················· | ···················· | ···················· | |
| E | 6994 | hCoV-19/Wales/PHWC-1655EB/2020\|EPI_ISL_484686\|2020 | ···················· | ···················· | ···················· | ···················· | ···················· | ···················· | |
| E | 6995 | hCoV-19/Wales/PHWC-165615/2020\|EPI_ISL_484689\|2020 | ···················· | ···················· | ···················· | ···················· | ···················· | ···················· | |
| E | 6996 | hCoV-19/Wales/PHWC-165633/2020\|EPI_ISL_484691\|2020 | ···················· | ···················· | ···················· | ···················· | ···················· | ···················· | |
| E | 6997 | hCoV-19/USA/LA-GBCL-26684475/2020\|EPI_ISL_485902\|2 | ···················· | ···················· | ···················· | ···················· | ···················· | ···················· | |
| E | 6998 | hCoV-19/USA/LA-GBCL-26684476/2020\|EPI_ISL_485903\|2 | ···················· | ···················· | ···················· | ···················· | ···················· | ···················· | |
| E | 6999 | hCoV-19/USA/LA-GBCL-26684504/2020\|EPI_ISL_485904\|2 | ···················· | ···················· | ···················· | ···················· | ···················· | ···················· | |
| E | 7000 | hCoV-19/USA/CT-UW-2427/2020\|EPI_ISL_485917\|2020-03 | ···················· | ···················A | ···················· | ···················· | ···················· | ···················· | 1 |
| E | 7001 | hCoV-19/USA/CT-UW-2432/2020\|EPI_ISL_485918\|2020-03 | ···················· | ···················· | ···················· | ···················· | ···················· | ···················· | |
| E | 7002 | hCoV-19/USA/CT-UW-2441/2020\|EPI_ISL_485919\|2020-03 | ···················· | ···················· | ···················· | ···················· | ···················· | ···················· | |
| E | 7003 | hCoV-19/USA/CT-UW-2462/2020\|EPI_ISL_485920\|2020-03 | ···················· | ···················· | ···················· | ···················· | ···················· | ···················· | |
| E | 7004 | hCoV-19/USA/CT-UW-2465/2020\|EPI_ISL_485921\|2020-03 | ···················· | ···················· | ···················· | ···················· | ···················· | ···················· | |
| E | 7005 | hCoV-19/USA/CT-UW-2469/2020\|EPI_ISL_485922\|2020-03 | ···················· | ···················· | ···················· | ···················· | ···················· | ···················· | |
| E | 7006 | hCoV-19/USA/CT-UW-2475/2020\|EPI_ISL_485923\|2020-03 | ···················· | ···················· | ···················· | ···················· | ···················· | ···················· | |
| E | 7007 | hCoV-19/USA/CT-UW-2481/2020\|EPI_ISL_485924\|2020-03 | ···················· | ···················A | ···················· | ···················· | ···················· | ···················· | 1 |
| E | 7008 | hCoV-19/USA/CT-UW-2488/2020\|EPI_ISL_485925\|2020-03 | ···················· | ···················· | ···················· | ···················· | ··T················· | ···················· | |
| E | 7009 | hCoV-19/USA/CT-UW-2671/2020\|EPI_ISL_485926\|2020-03 | ···················· | ···················· | ···················· | ···················· | ···················· | ···················· | |
| E | 7010 | hCoV-19/USA/CT-UW-2674/2020\|EPI_ISL_485927\|2020-03 | ···················· | ···················· | ···················· | ···················· | ···················· | ···················· | |
| E | 7011 | hCoV-19/USA/CT-UW-2676/2020\|EPI_ISL_485928\|2020-03 | ···················· | ···················· | ···················· | ···················· | ···················· | ···················· | |
| E | 7012 | hCoV-19/USA/CT-UW-3085/2020\|EPI_ISL_485929\|2020-03 | ···················· | ···················· | ···················· | ···················· | ···················· | ···················· | |
| E | 7013 | hCoV-19/USA/CT-UW-3094/2020\|EPI_ISL_485930\|2020-03 | ···················· | ···················· | ···················· | ···················· | ···················· | ···················· | |
| E | 7014 | hCoV-19/USA/CT-UW-3104/2020\|EPI_ISL_485932\|2020-03 | ···················· | ···················· | ···················· | ···················· | ···················· | ···················· | |
| E | 7015 | hCoV-19/USA/CT-UW-3106/2020\|EPI_ISL_485933\|2020-03 | ···················· | ···················· | ···················· | ···················· | ···················· | ···················· | |
| E | 7016 | hCoV-19/USA/CT-UW-3108/2020\|EPI_ISL_485934\|2020-03 | ···················· | ···················· | ···················· | ···················· | ···················· | ···················· | |
| E | 7017 | hCoV-19/USA/CT-UW-3118/2020\|EPI_ISL_485936\|2020-03 | ···················· | ···················· | ···················· | ···················· | ···················· | ···················· | |
| E | 7018 | hCoV-19/USA/CT-UW-3119/2020\|EPI_ISL_485937\|2020-03 | ···················· | ···················· | ···················· | ···················· | ···················· | ···················· | |
| E | 7019 | hCoV-19/USA/CT-UW-3120/2020\|EPI_ISL_485938\|2020-03 | ···················· | ···················· | ···················· | ···················· | ···················· | ···················· | |
| E | 7020 | hCoV-19/USA/CT-UW-3125/2020\|EPI_ISL_485940\|2020-03 | ···················· | ···················· | ···················· | ···················· | ···················· | ···················· | |
| E | 7021 | hCoV-19/USA/CT-UW-3126/2020\|EPI_ISL_485941\|2020-03 | ···················· | ···················· | ···················· | ···················· | ···················· | ···················· | |
| E | 7022 | hCoV-19/USA/CT-UW-3127/2020\|EPI_ISL_485942\|2020-03 | ···················· | ···················· | ···················· | ···················· | ···················· | ···················· | |
| E | 7023 | hCoV-19/USA/CT-UW-673/2020\|EPI_ISL_485944\|2020-03- | ···················· | ···················· | ···················· | ···················· | ···················· | ···················· | |
| E | 7024 | hCoV-19/USA/ID-UW-2041/2020\|EPI_ISL_485945\|2020-03 | ···················· | ···················· | ···················· | ···················· | ···················· | ···················· | |
| E | 7025 | hCoV-19/USA/ID-UW-2480/2020\|EPI_ISL_485946\|2020-03 | ···················· | ···················· | ···················· | ···················· | ···················· | ···················· | |
| E | 7026 | hCoV-19/USA/ID-UW-3036/2020\|EPI_ISL_485947\|2020-03 | ···················· | ···················· | ···················· | ···················· | ···················· | ···················· | |
| E | 7027 | hCoV-19/USA/OR-UW-679/2020\|EPI_ISL_485948\|2020-03- | ···················· | ···················· | ···················· | ···················· | ···················· | ···················· | |
| E | 7028 | hCoV-19/USA/UN-UW-2096/2020\|EPI_ISL_485950\|2020-04 | ···················· | ···················· | ···················· | ···················· | ···················· | ···················· | |
| E | 7029 | hCoV-19/USA/UN-UW-2099/2020\|EPI_ISL_485951\|2020-04 | ···················· | ···················· | ···················· | ···················· | ···················· | ···················· | |
| E | 7030 | hCoV-19/USA/UN-UW-2814/2020\|EPI_ISL_485952\|2020-04 | ···················· | ···················· | ···················· | ···················· | ···················· | ···················· | |
| E | 7031 | hCoV-19/USA/UN-UW-2820/2020\|EPI_ISL_485953\|2020-04 | ···················· | ···················· | ···················· | ···················· | ···················· | ···················· | |
| E | 7032 | hCoV-19/USA/UN-UW-2832/2020\|EPI_ISL_485954\|2020-04 | ···················· | ···················· | ···················· | ···················· | ···················· | ···················· | |
| E | 7033 | hCoV-19/USA/UN-UW-2834/2020\|EPI_ISL_485955\|2020-04 | ···················· | ···················· | ···················· | ···················· | ···················· | ···················· | |
| E | 7034 | hCoV-19/USA/UN-UW-2837/2020\|EPI_ISL_485956\|2020-04 | ···················· | ···················· | ···················· | ···················· | ···················· | ···················· | |
| E | 7035 | hCoV-19/USA/UN-UW-2849/2020\|EPI_ISL_485958\|2020-04 | ···················· | ···················· | ···················· | ···················· | ···················· | ···················· | |
| E | 7036 | hCoV-19/USA/UN-UW-2854/2020\|EPI_ISL_485960\|2020-04 | ···················· | ···················· | ···················· | ···················· | ···················· | ···················· | |
| E | 7037 | hCoV-19/USA/UN-UW-2855/2020\|EPI_ISL_485961\|2020-04 | ···················· | ···················· | ···················· | ···················· | ···················· | ···················· | |
| E | 7038 | hCoV-19/USA/UN-UW-2859/2020\|EPI_ISL_485963\|2020-04 | ···················· | ···················· | ···················· | ···················· | ···················· | ···················· | |
| E | 7039 | hCoV-19/USA/UN-UW-2863/2020\|EPI_ISL_485964\|2020-04 | ···················· | ···················· | ···················· | ···················· | ···················· | ···················· | |
| E | 7040 | hCoV-19/USA/UN-UW-2874/2020\|EPI_ISL_485967\|2020-04 | ···················· | ···················· | ···················· | ···················· | ···················· | ···················· | |
| E | 7041 | hCoV-19/USA/UN-UW-2890/2020\|EPI_ISL_485968\|2020-04 | ···················· | ···················· | ···················· | ···················· | ···················· | ···················· | |
| E | 7042 | hCoV-19/USA/UN-UW-2897/2020\|EPI_ISL_485969\|2020-04 | ···················· | ···················· | ···················· | ···················· | ···················· | ···················· | |
| E | 7043 | hCoV-19/USA/WA-UW-1764/2020\|EPI_ISL_485972\|2020-04 | ···················· | ···················· | ···················· | ···················· | ···················· | ···················· | |
| E | 7044 | hCoV-19/USA/WA-UW-1768/2020\|EPI_ISL_485973\|2020-04 | ···················· | ···················· | ···················· | ···················· | ···················· | ···················· | |
| E | 7045 | hCoV-19/USA/WA-UW-2020/2020\|EPI_ISL_485974\|2020-03 | ···················· | ···················· | ···················· | ···················· | ···················· | ···················· | |
| E | 7046 | hCoV-19/USA/WA-UW-2022/2020\|EPI_ISL_485975\|2020-03 | ···················· | ···················· | ···················· | ···················· | ···················· | ···················· | |
| E | 7047 | hCoV-19/USA/WA-UW-2029/2020\|EPI_ISL_485976\|2020-03 | ···················· | ···················· | ···················· | ···················· | ···················· | ···················· | |
| E | 7048 | hCoV-19/USA/WA-UW-2034/2020\|EPI_ISL_485977\|2020-03 | ···················· | ···················· | ···················· | ···················· | ···················· | ···················· | |
| E | 7049 | hCoV-19/USA/WA-UW-2053/2020\|EPI_ISL_485978\|2020-03 | ···················· | ···················· | ···················· | ···················· | ···················· | ···················· | |
| E | 7050 | hCoV-19/USA/WA-UW-2068/2020\|EPI_ISL_485979\|2020-03 | ···················· | ···················· | ···················· | ···················· | ···················· | ···················· | |
| E | 7051 | hCoV-19/USA/WA-UW-2071/2020\|EPI_ISL_485980\|2020-03 | ···················· | ···················· | ···················· | ···················· | ···················· | ···················· | |
| E | 7052 | hCoV-19/USA/WA-UW-2304/2020\|EPI_ISL_485981\|2020-03 | ···················· | ···················· | ···················· | ···················· | ···················· | ···················· | |
| E | 7053 | hCoV-19/USA/CA-CZB-1845/2020\|EPI_ISL_486158\|2020-0 | ···················· | ···················· | ···················· | ···················· | ···················· | ···················· | |
| E | 7054 | hCoV-19/USA/CA-CZB-1846/2020\|EPI_ISL_486159\|2020-0 | ···················· | ···················· | ···················· | ···················· | ···················· | ···················· | |
| E | 7055 | hCoV-19/USA/CA-CZB-1848/2020\|EPI_ISL_486160\|2020-0 | ···················· | ···················· | ···················· | ···················· | ···················· | ···················· | |
| E | 7056 | hCoV-19/USA/CA-CZB-1849/2020\|EPI_ISL_486161\|2020-0 | ···················· | ···················· | ···················· | ···················· | ···················· | ···················· | |
| E | 7057 | hCoV-19/USA/CA-CZB-1851/2020\|EPI_ISL_486162\|2020-0 | ···················· | ···················· | ···················· | ···················· | ···················· | ···················· | |
| E | 7058 | hCoV-19/USA/CA-CZB-1854/2020\|EPI_ISL_486163\|2020-0 | ···················· | ···················· | ···················· | ···················· | ···················· | ···················· | |
| E | 7059 | hCoV-19/USA/CA-CZB-1855/2020\|EPI_ISL_486164\|2020-0 | ···················· | ···················· | ···················· | ···················· | ······K············· | ···················· | |
| E | 7060 | hCoV-19/USA/CA-CZB-1857/2020\|EPI_ISL_486165\|2020-0 | ···················· | ···················· | ···················· | ···················· | ···················· | ···················· | |
| E | 7061 | hCoV-19/USA/CA-CZB-1859/2020\|EPI_ISL_486166\|2020-0 | ···················· | ···················· | ···················· | ···················· | ···················· | ···················· | |
| E | 7062 | hCoV-19/USA/CA-CZB-1861/2020\|EPI_ISL_486167\|2020-0 | ···················· | ···················· | ···················· | ···················· | ···················· | ···················· | |
| E | 7063 | hCoV-19/USA/CA-CZB-1863/2020\|EPI_ISL_486168\|2020-0 | ···················· | ···················· | ···················· | ···················· | ···················· | ···················· | |
| E | 7064 | hCoV-19/USA/CA-CZB-1871/2020\|EPI_ISL_486171\|2020-0 | ···················· | ···················· | ···················· | ···················· | ···················· | ···················· | |
| E | 7065 | hCoV-19/USA/CA-CZB-1872/2020\|EPI_ISL_486172\|2020-0 | ···················· | ···················· | ···················· | ···················· | ···················· | ···················· | |
| E | 7066 | hCoV-19/USA/CA-CZB-1873/2020\|EPI_ISL_486173\|2020-0 | ···················· | ···················· | ···················· | ···················· | ···················· | ···················· | |
| E | 7067 | hCoV-19/USA/CA-CZB-1874/2020\|EPI_ISL_486174\|2020-0 | ···················· | ···················· | ···················· | ···················· | ···················· | ···················· | |
| E | 7068 | hCoV-19/USA/CA-CZB-1877/2020\|EPI_ISL_486176\|2020-0 | ···················· | ···················· | ···················· | ···················· | ···················· | ···················· | |
| E | 7069 | hCoV-19/USA/CA-CZB-1878/2020\|EPI_ISL_486177\|2020-0 | ···················· | ···················· | ···················· | ···················· | ···················· | ···················· | |
| E | 7070 | hCoV-19/USA/CA-CZB-1879/2020\|EPI_ISL_486178\|2020-0 | ···················· | ···················· | ···················· | ···················· | ···················· | ···················· | |
| E | 7071 | hCoV-19/USA/CA-CZB-1880/2020\|EPI_ISL_486179\|2020-0 | ···················· | ···················· | ···················· | ···················· | ···················· | ···················· | |

| | | | | | | | | | |
|---|---|---|---|---|---|---|---|---|---|
| E | 7072 | hCoV-19/USA/CA-CZB-1881/2020\|EPI_ISL_486180\|2020-0 | .................... | .................... | .................... | .................... | .................... | .................... | |
| E | 7073 | hCoV-19/England/NORT-296C09/2020\|EPI_ISL_488138\|20 | .................... | .................... | .................... | .................... | .................... | .................... | |
| E | 7074 | hCoV-19/England/NORT-29ED1D/2020\|EPI_ISL_488139\|20 | .................... | .................... | .................... | .................... | .................... | .................... | |
| E | 7075 | hCoV-19/England/NORT-297A0E/2020\|EPI_ISL_488141\|20 | .................... | .A.................. | .................... | .................... | .................... | .................... | |
| E | 7076 | hCoV-19/England/NORT-2980ED/2020\|EPI_ISL_488142\|20 | .................... | .................... | .................... | .................... | .................... | .................... | |
| E | 7077 | hCoV-19/England/NORT-297154/2020\|EPI_ISL_488144\|20 | .................... | .................... | .................... | .................... | .................... | .................... | |
| E | 7078 | hCoV-19/England/NORT-296FE2/2020\|EPI_ISL_488145\|20 | .................... | .................... | .................... | .................... | .................... | .................... | |
| E | 7079 | hCoV-19/England/NORT-296BA2/2020\|EPI_ISL_488146\|20 | .................... | .................... | .................... | .................... | .................... | .................... | |
| E | 7080 | hCoV-19/England/NORT-29E4FA/2020\|EPI_ISL_488147\|20 | .................... | .................... | .................... | .................... | .................... | .................... | |
| E | 7081 | hCoV-19/England/NORT-29F4EA/2020\|EPI_ISL_488150\|20 | .................... | .................... | .................... | .................... | .................... | .................... | |
| E | 7082 | hCoV-19/England/NORT-297AD1/2020\|EPI_ISL_488151\|20 | .................... | .................... | .................... | .................... | .................... | .................... | |
| E | 7083 | hCoV-19/England/NORT-296E30/2020\|EPI_ISL_488152\|20 | .................... | .................... | .................... | .................... | .................... | .................... | |
| E | 7084 | hCoV-19/England/NORT-297F2D/2020\|EPI_ISL_488153\|20 | .................... | .................... | .................... | .................... | .................... | .................... | |
| E | 7085 | hCoV-19/England/NORT-297BA1/2020\|EPI_ISL_488154\|20 | .................... | .................... | .................... | .................... | .................... | .................... | |
| E | 7086 | hCoV-19/England/NORT-29C6AB/2020\|EPI_ISL_488156\|20 | .................... | .................... | .................... | .................... | .................... | .................... | |
| E | 7087 | hCoV-19/England/NORT-29D0BB/2020\|EPI_ISL_488157\|20 | .................... | .................... | .................... | .................... | .................... | .................... | |
| E | 7088 | hCoV-19/England/NORT-29D886/2020\|EPI_ISL_488158\|20 | .................... | .................... | .................... | .................... | .................... | .................... | |
| E | 7089 | hCoV-19/England/NORT-297181/2020\|EPI_ISL_488159\|20 | .................... | .................... | .................... | .................... | .................... | .................... | |
| E | 7090 | hCoV-19/England/NORT-29D446/2020\|EPI_ISL_488160\|20 | .................... | .................... | .................... | .................... | .................... | .................... | |
| E | 7091 | hCoV-19/England/NORT-296DCA/2020\|EPI_ISL_488161\|20 | .................... | .................... | .................... | .................... | .................... | .................... | |
| E | 7092 | hCoV-19/England/NORT-29E24B/2020\|EPI_ISL_488162\|20 | .................... | .................... | .................... | .................... | .................... | .................... | |
| E | 7093 | hCoV-19/England/NORT-29D4BF/2020\|EPI_ISL_488163\|20 | .................... | .................... | .................... | .................... | .................... | .................... | |
| E | 7094 | hCoV-19/England/NORT-29F57E/2020\|EPI_ISL_488164\|20 | .................... | .................... | .................... | .................... | .................... | .................... | |
| E | 7095 | hCoV-19/England/NORT-29C9DF/2020\|EPI_ISL_488166\|20 | .................... | .................... | .................... | .................... | .................... | .................... | |
| E | 7096 | hCoV-19/England/NORT-296DF7/2020\|EPI_ISL_488168\|20 | .................... | .................... | .................... | .................... | .................... | .................... | |
| E | 7097 | hCoV-19/England/NORT-29E22D/2020\|EPI_ISL_488171\|20 | .................... | .................... | .................... | .................... | .................... | .................... | |
| E | 7098 | hCoV-19/England/NORT-29E8A3/2020\|EPI_ISL_488172\|20 | .................... | .................... | .................... | .................... | .................... | .................... | |
| E | 7099 | hCoV-19/England/NORT-297145/2020\|EPI_ISL_488174\|20 | .................... | .................... | .................... | .................... | .................... | .................... | |
| E | 7100 | hCoV-19/England/NORT-29E9EC/2020\|EPI_ISL_488176\|20 | .................... | .................... | .................... | .................... | .................... | .................... | |
| E | 7101 | hCoV-19/England/NORT-292FAA/2020\|EPI_ISL_488179\|20 | .................... | .................... | .................... | .................... | .................... | .................... | |
| E | 7102 | hCoV-19/England/NORT-292F9B/2020\|EPI_ISL_488180\|20 | .................... | .................... | .................... | .................... | .................... | .................... | |
| E | 7103 | hCoV-19/England/NORT-292F22/2020\|EPI_ISL_488181\|20 | .................... | .................... | .................... | .................... | .................... | .................... | |
| E | 7104 | hCoV-19/England/NORT-2930C4/2020\|EPI_ISL_488182\|20 | .................... | .................... | .................... | .................... | .................... | .................... | |
| E | 7105 | hCoV-19/England/NORT-292EF8/2020\|EPI_ISL_488183\|20 | .................... | .................... | .................... | .................... | .................... | .................... | |
| E | 7106 | hCoV-19/England/NORT-292FE6/2020\|EPI_ISL_488184\|20 | .................... | .................... | .................... | .................... | .................... | .................... | |
| E | 7107 | hCoV-19/England/NORT-29306A/2020\|EPI_ISL_488186\|20 | .................... | .................... | .................... | .................... | .................... | .................... | |
| E | 7108 | hCoV-19/England/BRIS-12F9F0/2020\|EPI_ISL_488187\|20 | .................... | .................... | .................... | .................... | .................... | .................... | |
| E | 7109 | hCoV-19/England/BRIS-12E636/2020\|EPI_ISL_488188\|20 | .................... | .............A...... | .................... | .................... | .................... | .................... | |
| E | 7110 | hCoV-19/England/BRIS-12F9B4/2020\|EPI_ISL_488189\|20 | .................... | .................... | .................... | .................... | .................... | .................... | |
| E | 7111 | hCoV-19/England/BRIS-12F76F/2020\|EPI_ISL_488190\|20 | .................... | .................... | .................... | .................... | .................... | .................... | |
| E | 7112 | hCoV-19/England/BRIS-12ED22/2020\|EPI_ISL_488191\|20 | .................... | .................... | .................... | .................... | .................... | .................... | |
| E | 7113 | hCoV-19/England/BRIS-12F9E1/2020\|EPI_ISL_488192\|20 | .................... | .................... | .................... | .................... | .................... | .................... | |
| E | 7114 | hCoV-19/England/BRIS-12FA39/2020\|EPI_ISL_488193\|20 | .................... | .................... | .................... | .................... | .................... | .................... | |
| E | 7115 | hCoV-19/England/BRIS-12F51A/2020\|EPI_ISL_488194\|20 | .................... | .................... | .................... | .................... | .................... | .................... | |
| E | 7116 | hCoV-19/England/BRIS-12FB36/2020\|EPI_ISL_488196\|20 | .................... | .................... | .................... | .................... | .................... | .................... | |
| E | 7117 | hCoV-19/England/BRIS-12F091/2020\|EPI_ISL_488197\|20 | .................... | .................... | .................... | .................... | .................... | .................... | |
| E | 7118 | hCoV-19/England/BRIS-12F79C/2020\|EPI_ISL_488199\|20 | .................... | .................... | .................... | .................... | .................... | .................... | |
| E | 7119 | hCoV-19/England/CAMB-1ACB9B/2020\|EPI_ISL_489469\|20 | .................... | .................... | .................... | .................... | .................... | .................... | |
| E | 7120 | hCoV-19/Scotland/GCVR-171A83/2020\|EPI_ISL_489470\|2 | .................... | .................... | .................... | .................... | .................... | .................... | |
| E | 7121 | hCoV-19/England/CAMB-1ACA61/2020\|EPI_ISL_489472\|20 | .................... | .................... | .................... | .................... | .................... | .................... | |
| E | 7122 | hCoV-19/England/CAMB-1AC8D0/2020\|EPI_ISL_489473\|20 | .................... | .................... | .................... | .................... | .................... | .................... | |
| E | 7123 | hCoV-19/England/CAMB-1ACC2F/2020\|EPI_ISL_489475\|20 | .................... | .................... | .................... | .................... | .................... | .................... | |
| E | 7124 | hCoV-19/England/CAMB-1ACCA7/2020\|EPI_ISL_489477\|20 | .................... | .................... | .................... | .................... | .................... | .................... | |
| E | 7125 | hCoV-19/England/CAMB-1AC75B/2020\|EPI_ISL_489478\|20 | .................... | .................... | .................... | .................... | .................... | .................... | |
| E | 7126 | hCoV-19/England/CAMB-1ACABC/2020\|EPI_ISL_489479\|20 | .................... | .................... | .................... | .................... | .................... | .................... | |
| E | 7127 | hCoV-19/Scotland/GCVR-171A56/2020\|EPI_ISL_489480\|2 | .................... | ...................A | .................... | .................... | .................... | .................... | 1 |
| E | 7128 | hCoV-19/England/CAMB-1AC5CA/2020\|EPI_ISL_489481\|20 | .................... | .................... | .................... | .................... | .................... | .................... | |
| E | 7129 | hCoV-19/England/CAMB-1ACEB0/2020\|EPI_ISL_489482\|20 | .................... | .................... | .................... | .................... | .................... | .................... | |
| E | 7130 | hCoV-19/England/CAMB-1AC418/2020\|EPI_ISL_489483\|20 | .................... | .................... | .................... | .................... | .................... | .................... | |
| E | 7131 | hCoV-19/England/CAMB-1AC6B8/2020\|EPI_ISL_489484\|20 | .................... | .................... | .................... | .................... | .................... | .................... | |
| E | 7132 | hCoV-19/England/CAMB-1ACF62/2020\|EPI_ISL_489485\|20 | .................... | .................... | .................... | .................... | .................... | .................... | |
| E | 7133 | hCoV-19/England/CAMB-1AC885/2020\|EPI_ISL_489486\|20 | .................... | .................... | .................... | .................... | .................... | .................... | |
| E | 7134 | hCoV-19/England/CAMB-1AD417/2020\|EPI_ISL_489487\|20 | .................... | .................... | .................... | .................... | .................... | .................... | |
| E | 7135 | hCoV-19/England/CAMB-1AD383/2020\|EPI_ISL_489488\|20 | .................... | .................... | .................... | .................... | .................... | .................... | |
| E | 7136 | hCoV-19/Scotland/GCVR-171968/2020\|EPI_ISL_489489\|2 | .................... | .................... | .................... | .................... | .................... | .................... | |
| E | 7137 | hCoV-19/England/CAMB-1AC779/2020\|EPI_ISL_489490\|20 | .................... | .A.................. | .................... | .................... | .................... | .................... | |
| E | 7138 | hCoV-19/England/CAMB-1AD3B0/2020\|EPI_ISL_489492\|20 | .................... | .................... | .................... | .................... | .................... | .................... | |
| E | 7139 | hCoV-19/England/CAMB-1ACE0B/2020\|EPI_ISL_489493\|20 | .................... | .................... | .................... | .................... | .................... | .................... | |
| E | 7140 | hCoV-19/England/CAMB-1AD356/2020\|EPI_ISL_489494\|20 | .................... | ..........A......... | .................... | .................... | .................... | .................... | |
| E | 7141 | hCoV-19/England/CAMB-1AD392/2020\|EPI_ISL_489496\|20 | .................... | .................... | .................... | .................... | .................... | .................... | |
| E | 7142 | hCoV-19/England/CAMB-1AC66D/2020\|EPI_ISL_489497\|20 | .................... | .................... | .................... | .................... | .................... | .................... | |
| E | 7143 | hCoV-19/England/CAMB-1AC560/2020\|EPI_ISL_489498\|20 | .................... | .................... | .................... | .................... | .................... | .................... | |
| E | 7144 | hCoV-19/England/CAMB-1AD347/2020\|EPI_ISL_489499\|20 | .................... | ..........A......... | .................... | .................... | .................... | .................... | |
| E | 7145 | hCoV-19/England/NORT-2948F8/2020\|EPI_ISL_490451\|20 | .................... | .A.................. | .................... | .................... | .................... | .................... | |
| E | 7146 | hCoV-19/England/NORT-294904/2020\|EPI_ISL_490452\|20 | .................... | .................... | .................... | .................... | .................... | .................... | |
| E | 7147 | hCoV-19/England/NORT-294913/2020\|EPI_ISL_490453\|20 | .................... | .................... | .................... | .................... | .................... | .................... | |
| E | 7148 | hCoV-19/England/NORT-294B1D/2020\|EPI_ISL_490461\|20 | .................... | .................... | .................... | .................... | .................... | .................... | |
| E | 7149 | hCoV-19/England/NORT-294B86/2020\|EPI_ISL_490467\|20 | .................... | .................... | .................... | .................... | .................... | .................... | |
| E | 7150 | hCoV-19/England/NORT-29EA07/2020\|EPI_ISL_490469\|20 | .................... | .................... | .................... | .................... | .................... | .................... | |
| E | 7151 | hCoV-19/England/NORT-29EAAD/2020\|EPI_ISL_490472\|20 | .................... | .................... | .................... | .................... | .................... | .................... | |

| | | | | | | | |
|---|---|---|---|---|---|---|---|
| E | 7152 hCoV-19/England/NORT-29EAE9/2020\|EPI_ISL_490473\|20 | .................... | .................... | .................... | .................... | .................... | .................... |
| E | 7153 hCoV-19/England/NORT-29EB04/2020\|EPI_ISL_490474\|20 | .................... | .................... | .................... | .................... | .................... | .................... |
| E | 7154 hCoV-19/England/NORT-29EC6B/2020\|EPI_ISL_490478\|20 | .................... | .................... | .................... | .................... | .................... | .................... |
| E | 7155 hCoV-19/England/NORT-29EC7A/2020\|EPI_ISL_490479\|20 | .................... | .................... | .................... | .................... | .................... | .................... |
| E | 7156 hCoV-19/England/NORT-29ECA7/2020\|EPI_ISL_490481\|20 | .................... | .................... | .................... | .................... | .................... | .................... |
| E | 7157 hCoV-19/England/NORT-29ECF2/2020\|EPI_ISL_490482\|20 | .................... | .................... | .................... | .................... | .................... | .................... |
| E | 7158 hCoV-19/England/NORT-29EDD1/2020\|EPI_ISL_490485\|20 | .................... | .................... | .................... | .................... | .................... | .................... |
| E | 7159 hCoV-19/England/NORT-29EF26/2020\|EPI_ISL_490487\|20 | .................... | .................... | .................... | .................... | .................... | .................... |
| E | 7160 hCoV-19/England/NORT-29F5F6/2020\|EPI_ISL_490492\|20 | .................... | .................... | .................... | .................... | .................... | .................... |
| E | 7162 hCoV-19/USA/VA-DCLS-0649/2020\|EPI_ISL_495321\|2020- | .................... | .................... | .................... | .................... | .................... | .................... |
| E | 7163 hCoV-19/USA/VA-DCLS-0650/2020\|EPI_ISL_495322\|2020- | .................... | .................... | .................... | .................... | .................... | .................... |
| E | 7164 hCoV-19/USA/VA-DCLS-0653/2020\|EPI_ISL_495325\|2020- | .................... | .................... | .................... | .................... | .................... | .................... |
| E | 7165 hCoV-19/USA/VA-DCLS-0654/2020\|EPI_ISL_495326\|2020- | .................... | .................... | .................... | .................... | .................... | .................... |
| E | 7166 hCoV-19/USA/VA-DCLS-0656/2020\|EPI_ISL_495327\|2020- | .................... | .................... | .................... | .................... | .................... | .................... |
| E | 7167 hCoV-19/USA/VA-DCLS-0657/2020\|EPI_ISL_495328\|2020- | .................... | .................... | .................... | .................... | .................... | .................... |
| E | 7168 hCoV-19/USA/VA-DCLS-0658/2020\|EPI_ISL_495329\|2020- | .................... | .................... | .................... | .................... | .................... | .................... |
| E | 7169 hCoV-19/Japan/NGS-CD-365/2020\|EPI_ISL_481263\|2020- | .................... | .................... | .................... | .................... | .................... | .................... |
| E | 7170 hCoV-19/USA/MD-MDH-0016/2020\|EPI_ISL_481265\|2020-0 | .................... | .................... | .................... | .................... | .................... | .................... |
| E | 7171 hCoV-19/USA/MD-MDH-0017/2020\|EPI_ISL_481266\|2020-0 | .................... | .................... | .................... | .................... | .................... | .................... |
| E | 7172 hCoV-19/USA/MD-MDH-0019/2020\|EPI_ISL_481267\|2020-0 | .................... | .................... | .................... | .................... | .................... | .................... |
| E | 7173 hCoV-19/USA/MD-MDH-0020/2020\|EPI_ISL_481268\|2020-0 | .................... | .................... | .................... | .................... | .................... | .................... |
| E | 7174 hCoV-19/USA/MD-MDH-0021/2020\|EPI_ISL_481269\|2020-0 | .................... | .................... | .................... | .................... | .................... | .................... |
| E | 7175 hCoV-19/USA/MD-MDH-0023/2020\|EPI_ISL_481270\|2020-0 | .................... | .................... | .................... | .................... | .................... | .................... |
| E | 7176 hCoV-19/USA/MD-MDH-0025/2020\|EPI_ISL_481271\|2020-0 | .................... | .................... | .................... | .................... | .................... | .................... |
| E | 7177 hCoV-19/USA/MD-MDH-0026/2020\|EPI_ISL_481272\|2020-0 | .................... | .................... | .................... | .................... | .................... | .................... |
| E | 7178 hCoV-19/USA/MD-MDH-0027/2020\|EPI_ISL_481273\|2020-0 | .................... | .................... | .................... | .................... | ......T............. | .................... |
| E | 7179 hCoV-19/USA/MD-MDH-0028/2020\|EPI_ISL_481274\|2020-0 | .................... | .................... | .................... | .................... | ......T............. | .................... |
| E | 7180 hCoV-19/USA/MD-MDH-0031/2020\|EPI_ISL_481276\|2020-0 | .................... | .................... | .................... | .................... | .................... | .................... |
| E | 7181 hCoV-19/USA/MD-MDH-0032/2020\|EPI_ISL_481277\|2020-0 | .................... | .................... | .................... | .................... | .................... | .................... |
| E | 7182 hCoV-19/USA/MD-MDH-0035/2020\|EPI_ISL_481280\|2020-0 | .................... | .................... | .................... | .................... | .................... | .................... |
| E | 7183 hCoV-19/USA/MD-MDH-0036/2020\|EPI_ISL_481281\|2020-0 | .................... | .................... | .................... | .................... | .................... | .................... |
| E | 7184 hCoV-19/USA/MD-MDH-0039/2020\|EPI_ISL_481282\|2020-0 | .................... | .................... | .................... | .................... | .................... | .................... |
| E | 7185 hCoV-19/USA/NY-NYU-VC-003/2020\|EPI_ISL_481283\|2020 | .................... | .................... | .................... | .................... | .................... | .................... |
| E | 7186 hCoV-19/England/BIRM-608D9/2020\|EPI_ISL_483901\|202 | .................... | .................... | .................... | .................... | .................... | .................... |
| E | 7187 hCoV-19/England/BIRM-60921/2020\|EPI_ISL_483905\|202 | .................... | .................... | .................... | .................... | .................... | .................... |
| E | 7188 hCoV-19/England/BIRM-60930/2020\|EPI_ISL_483906\|202 | .................... | .................... | .................... | .................... | .................... | .................... |
| E | 7189 hCoV-19/England/BIRM-6096D/2020\|EPI_ISL_483909\|202 | .................... | .................... | .................... | .................... | .................... | .................... |
| E | 7190 hCoV-19/England/BIRM-6098B/2020\|EPI_ISL_483911\|202 | .................... | .................... | .................... | .................... | .................... | .................... |
| E | 7191 hCoV-19/England/CAMB-1B5124/2020\|EPI_ISL_483913\|20 | .................... | .................... | .................... | .................... | .................... | .................... |
| E | 7192 hCoV-19/England/CAMB-1B51D9/2020\|EPI_ISL_483914\|20 | .................... | .................... | .................... | .................... | .................... | .................... |
| E | 7193 hCoV-19/England/CAMB-1B51F7/2020\|EPI_ISL_483915\|20 | .................... | .................... | .................... | .................... | .................... | .................... |
| E | 7194 hCoV-19/USA/WA-UW-5388/2020\|EPI_ISL_486102\|2020-04 | .................... | .................... | .................... | .................... | .................... | .................... |
| E | 7195 hCoV-19/USA/WA-UW-5429/2020\|EPI_ISL_486103\|2020-03 | .................... | .................... | .................... | .................... | .................... | .................... |
| E | 7196 hCoV-19/USA/WA-UW-682/2020\|EPI_ISL_486105\|2020-03- | .................... | .................... | .................... | .................... | .................... | .................... |
| E | 7197 hCoV-19/USA/WA-UW-688/2020\|EPI_ISL_486106\|2020-03- | .................... | .................... | .................... | .................... | .................... | .................... |
| E | 7198 hCoV-19/USA/WA-UW-7166/2020\|EPI_ISL_486108\|2020-04 | .................... | .................... | .................... | .................... | .................... | .................... |
| E | 7199 hCoV-19/USA/WA-UW-7178/2020\|EPI_ISL_486109\|2020-04 | .................... | .................... | .................... | .................... | .................... | .................... |
| E | 7200 hCoV-19/USA/WA-UW-7399/2020\|EPI_ISL_486110\|2020-04 | .................... | .................... | .................... | .................... | .................... | .................... |
| E | 7201 hCoV-19/USA/WA-UW-8150/2020\|EPI_ISL_486112\|2020-04 | .................... | .................... | .................... | .................... | .................... | .................... |
| E | 7202 hCoV-19/USA/WA-UW-822/2020\|EPI_ISL_486113\|2020-03- | .................... | .................... | .................... | .................... | .................... | .................... |
| E | 7203 hCoV-19/USA/WA-UW-909/2020\|EPI_ISL_486114\|2020-03- | .................... | .................... | .................... | .................... | .................... | .................... |
| E | 7204 hCoV-19/USA/CA-CZB-1778/2020\|EPI_ISL_486115\|2020-0 | .................... | .................... | .................... | .................... | .................... | .................... |
| E | 7205 hCoV-19/USA/CA-CZB-1782/2020\|EPI_ISL_486116\|2020-0 | .................... | .................... | .................... | .................... | .................... | .................... |
| E | 7206 hCoV-19/USA/CA-CZB-1783/2020\|EPI_ISL_486117\|2020-0 | .................... | .A.................. | .................... | .................... | .................... | .................... |
| E | 7207 hCoV-19/USA/CA-CZB-1785/2020\|EPI_ISL_486118\|2020-0 | .................... | .................... | .................... | .................... | .................... | .................... |
| E | 7208 hCoV-19/USA/CA-CZB-1790/2020\|EPI_ISL_486121\|2020-0 | .................... | .................... | .................... | .................... | .................... | .................... |
| E | 7209 hCoV-19/USA/CA-CZB-1793/2020\|EPI_ISL_486122\|2020-0 | .................... | .................... | .................... | .................... | .................... | .................... |
| E | 7210 hCoV-19/USA/CA-CZB-1794/2020\|EPI_ISL_486123\|2020-0 | .................... | .................... | .................... | .................... | .................... | .................... |
| E | 7211 hCoV-19/USA/CA-CZB-1795/2020\|EPI_ISL_486124\|2020-0 | .................... | .................... | .................... | .................... | .................... | .................... |
| E | 7212 hCoV-19/USA/CA-CZB-1796/2020\|EPI_ISL_486125\|2020-0 | .................... | .................... | .................... | .................... | .................... | .................... |
| E | 7213 hCoV-19/USA/CA-CZB-1797/2020\|EPI_ISL_486126\|2020-0 | .................... | .................... | .................... | .................... | .................... | .................... |
| E | 7214 hCoV-19/USA/CA-CZB-1798/2020\|EPI_ISL_486127\|2020-0 | .................... | .................... | .................... | .................... | .................... | .................... |
| E | 7215 hCoV-19/USA/CA-CZB-1799/2020\|EPI_ISL_486128\|2020-0 | .................... | .................... | .................... | .................... | .................... | .................... |
| E | 7216 hCoV-19/USA/CA-CZB-1800/2020\|EPI_ISL_486129\|2020-0 | .................... | .................... | .................... | .................... | .................... | .................... |
| E | 7217 hCoV-19/USA/CA-CZB-1801/2020\|EPI_ISL_486130\|2020-0 | .................... | .................... | .................... | .................... | .................... | .................... |
| E | 7218 hCoV-19/USA/CA-CZB-1802/2020\|EPI_ISL_486131\|2020-0 | .................... | .................... | .................... | .................... | .................... | .................... |
| E | 7219 hCoV-19/USA/CA-CZB-1803/2020\|EPI_ISL_486132\|2020-0 | .................... | .................... | .................... | .................... | .................... | .................... |
| E | 7220 hCoV-19/USA/CA-CZB-1817/2020\|EPI_ISL_486134\|2020-0 | .................... | .A.................. | .................... | .................... | .................... | .................... |
| E | 7221 hCoV-19/USA/CA-CZB-1820/2020\|EPI_ISL_486137\|2020-0 | .................... | .................... | .................... | .................... | .................... | .................... |
| E | 7222 hCoV-19/USA/CA-CZB-1822/2020\|EPI_ISL_486138\|2020-0 | .................... | .................... | .................... | .................... | .................... | .................... |
| E | 7223 hCoV-19/USA/CA-CZB-1823/2020\|EPI_ISL_486139\|2020-0 | .................... | .................... | .................... | .................... | .................... | .................... |
| E | 7224 hCoV-19/USA/CA-CZB-1825/2020\|EPI_ISL_486141\|2020-0 | .................... | .................... | .................... | .................... | .................... | .................... |
| E | 7225 hCoV-19/USA/CA-CZB-1826/2020\|EPI_ISL_486142\|2020-0 | .................... | .................... | .................... | .................... | .................... | .................... |
| E | 7226 hCoV-19/USA/CA-CZB-1827/2020\|EPI_ISL_486143\|2020-0 | .................... | .................... | .................... | .................... | .................... | .................... |
| E | 7227 hCoV-19/USA/CA-CZB-1828/2020\|EPI_ISL_486144\|2020-0 | .................... | .................... | .................... | .................... | .................... | .................... |
| E | 7228 hCoV-19/USA/CA-CZB-1829/2020\|EPI_ISL_486145\|2020-0 | .................... | .................... | .................... | .................... | .................... | .................... |
| E | 7229 hCoV-19/USA/CA-CZB-1830/2020\|EPI_ISL_486146\|2020-0 | .................... | .................... | .................... | .................... | .................... | .................... |
| E | 7230 hCoV-19/USA/CA-CZB-1832/2020\|EPI_ISL_486147\|2020-0 | .................... | .................... | .................... | .................... | .................... | .................... |
| E | 7231 hCoV-19/USA/CA-CZB-1833/2020\|EPI_ISL_486148\|2020-0 | .................... | .................... | .................... | .................... | .................... | .................... |
| E | 7232 hCoV-19/USA/CA-CZB-1835/2020\|EPI_ISL_486149\|2020-0 | .................... | .................... | .................... | .................... | .................... | .................... |

| | | | | | | | |
|---|---|---|---|---|---|---|---|
| E | 7233 hCoV-19/USA/CA-CZB-1836/2020\|EPI_ISL_486150\|2020-0 | .................... | .................... | .................... | .................... | .................... | .................... |
| E | 7234 hCoV-19/USA/CA-CZB-1838/2020\|EPI_ISL_486151\|2020-0 | .................... | .................... | .................... | .................... | .................... | .................... |
| E | 7235 hCoV-19/USA/CA-CZB-1840/2020\|EPI_ISL_486153\|2020-0 | .................... | .................... | .................... | .................... | .................... | .................... |
| E | 7236 hCoV-19/USA/CA-CZB-1841/2020\|EPI_ISL_486154\|2020-0 | .................... | .................... | .................... | .................... | .................... | .................... |
| E | 7237 hCoV-19/USA/CA-CZB-1842/2020\|EPI_ISL_486155\|2020-0 | .................... | .................... | .................... | .................... | .................... | .................... |
| E | 7238 hCoV-19/USA/CA-CZB-1843/2020\|EPI_ISL_486156\|2020-0 | .................... | .................... | .................... | .................... | .................... | .................... |
| E | 7239 hCoV-19/USA/CA-CZB-1844/2020\|EPI_ISL_486157\|2020-0 | .................... | .................... | .................... | .................... | .................... | .................... |
| E | 7240 hCoV-19/USA/CA-CZB-1882/2020\|EPI_ISL_486181\|2020-0 | .................... | .................... | .................... | .................... | .................... | .................... |
| E | 7241 hCoV-19/USA/CA-CZB-1883/2020\|EPI_ISL_486182\|2020-0 | .................... | .................... | .................... | .................... | .................... | .................... |
| E | 7242 hCoV-19/USA/CA-CZB-1884/2020\|EPI_ISL_486183\|2020-0 | .................... | .................... | .................... | .................... | .................... | .................... |
| E | 7243 hCoV-19/USA/CA-CZB-1885/2020\|EPI_ISL_486184\|2020-0 | .................... | .................... | .................... | .................... | .................... | .................... |
| E | 7244 hCoV-19/USA/CA-CZB-1886/2020\|EPI_ISL_486185\|2020-0 | .................... | .................... | .................... | .................... | .................... | .................... |
| E | 7245 hCoV-19/USA/CA-CZB-1887/2020\|EPI_ISL_486186\|2020-0 | .................... | .................... | .................... | .................... | .................... | .................... |
| E | 7246 hCoV-19/USA/CA-CZB-1888/2020\|EPI_ISL_486187\|2020-0 | .................... | .................... | .................... | .................... | .................... | .................... |
| E | 7247 hCoV-19/USA/CA-CZB-1889/2020\|EPI_ISL_486188\|2020-0 | .................... | .................... | .................... | .................... | .................... | .................... |
| E | 7248 hCoV-19/USA/CA-CZB-1890/2020\|EPI_ISL_486189\|2020-0 | .................... | .................... | .................... | .................... | .................... | .................... |
| E | 7249 hCoV-19/USA/CA-CZB-1892/2020\|EPI_ISL_486191\|2020-0 | .................... | .................... | .................... | .................... | .................... | .................... |
| E | 7250 hCoV-19/USA/CA-CZB-1893/2020\|EPI_ISL_486192\|2020-0 | .................... | .................... | .................... | .................... | .................... | .................... |
| E | 7251 hCoV-19/USA/CA-CZB-1894/2020\|EPI_ISL_486193\|2020-0 | .................... | .................... | .................... | .................... | .................... | .................... |
| E | 7252 hCoV-19/USA/CA-CZB-1895/2020\|EPI_ISL_486194\|2020-0 | .................... | .................... | .................... | .................... | .................... | .................... |
| E | 7253 hCoV-19/USA/CA-CZB-1896/2020\|EPI_ISL_486195\|2020-0 | .................... | .................... | .................... | .................... | .................... | .................... |
| E | 7254 hCoV-19/USA/CA-CZB-1897/2020\|EPI_ISL_486196\|2020-0 | .................... | .................... | .................... | .................... | .................... | .................... |
| E | 7255 hCoV-19/USA/CA-CZB-1899/2020\|EPI_ISL_486198\|2020-0 | .................... | .................... | .................... | .................... | .................... | .................... |
| E | 7256 hCoV-19/Germany/NW-MPP-12/2020\|EPI_ISL_487400\|2020 | .................... | .................... | .................... | .................... | .................... | .................... |
| E | 7257 hCoV-19/Germany/NW-MPP-13/2020\|EPI_ISL_487401\|2020 | .................... | .................... | .................... | .................... | .................... | .................... |
| E | 7258 hCoV-19/Germany/NW-MPP-14/2020\|EPI_ISL_487402\|2020 | .................... | .................... | .................... | .................... | .................... | .................... |
| E | 7259 hCoV-19/Germany/NW-MPP-15/2020\|EPI_ISL_487403\|2020 | .................... | .................... | .................... | .................... | .................... | .................... |
| E | 7260 hCoV-19/Germany/NW-MPP-16/2020\|EPI_ISL_487404\|2020 | .................... | .................... | .................... | .................... | .................... | .................... |
| E | 7261 hCoV-19/Germany/NW-MPP-17/2020\|EPI_ISL_487405\|2020 | .................... | .................... | .................... | .................... | .................... | .................... |
| E | 7262 hCoV-19/Germany/NW-MPP-18/2020\|EPI_ISL_487406\|2020 | .................... | .................... | .................... | .................... | .................... | .................... |
| E | 7263 hCoV-19/Germany/NW-MPP-19/2020\|EPI_ISL_487407\|2020 | .................... | .................... | .................... | .................... | .................... | .................... |
| E | 7264 hCoV-19/Germany/NW-MPP-2/2020\|EPI_ISL_487408\|2020- | .................... | .................... | .................... | .................... | .................... | .................... |
| E | 7265 hCoV-19/Germany/NW-MPP-20/2020\|EPI_ISL_487409\|2020 | .................... | .................... | .................... | .................... | .................... | .................... |
| E | 7266 hCoV-19/Germany/NW-MPP-21/2020\|EPI_ISL_487410\|2020 | .................... | .................... | .................... | .................... | .................... | .................... |
| E | 7267 hCoV-19/Germany/NW-MPP-23/2020\|EPI_ISL_487412\|2020 | .................... | .................... | .................... | .................... | .................... | .................... |
| E | 7268 hCoV-19/Germany/NW-MPP-24/2020\|EPI_ISL_487413\|2020 | .................... | .................... | .................... | .................... | .................... | .................... |
| E | 7269 hCoV-19/Germany/NW-MPP-25/2020\|EPI_ISL_487414\|2020 | .................... | .................... | .................... | .................... | .................... | .................... |
| E | 7270 hCoV-19/Germany/NW-MPP-27/2020\|EPI_ISL_487416\|2020 | .................... | .................... | .................... | .................... | .................... | .................... |
| E | 7271 hCoV-19/Germany/NW-MPP-28/2020\|EPI_ISL_487417\|2020 | .................... | .................... | .................... | .................... | .................... | .................... |
| E | 7272 hCoV-19/Germany/NW-MPP-29/2020\|EPI_ISL_487418\|2020 | .................... | .................... | .................... | .................... | .................... | .................... |
| E | 7273 hCoV-19/Germany/NW-MPP-3/2020\|EPI_ISL_487419\|2020- | .................... | .................... | .................... | .................... | .................... | .................... |
| E | 7274 hCoV-19/Germany/NW-MPP-30/2020\|EPI_ISL_487420\|2020 | .................... | .................... | .................... | .................... | .................... | .................... |
| E | 7275 hCoV-19/Germany/NW-MPP-31/2020\|EPI_ISL_487421\|2020 | .................... | .................... | .................... | .................... | .................... | .................... |
| E | 7276 hCoV-19/Germany/NW-MPP-32/2020\|EPI_ISL_487422\|2020 | .................... | .................... | .................... | .................... | .................... | .................... |
| E | 7277 hCoV-19/Germany/NW-MPP-33/2020\|EPI_ISL_487423\|2020 | .................... | .................... | .................... | .................... | .................... | .................... |
| E | 7278 hCoV-19/Germany/NW-MPP-34/2020\|EPI_ISL_487424\|2020 | .................... | .................... | .................... | .................... | .................... | .................... |
| E | 7279 hCoV-19/Germany/NW-MPP-35/2020\|EPI_ISL_487425\|2020 | .................... | .................... | .................... | .................... | .................... | .................... |
| E | 7280 hCoV-19/Germany/NW-MPP-4/2020\|EPI_ISL_487426\|2020- | .................... | .................... | .................... | .................... | .................... | .................... |
| E | 7281 hCoV-19/Germany/NW-MPP-5/2020\|EPI_ISL_487427\|2020- | .................... | .................... | .................... | .................... | .................... | .................... |
| E | 7282 hCoV-19/Germany/NW-MPP-7/2020\|EPI_ISL_487429\|2020- | .................... | .................... | .................... | .................... | .................... | .................... |
| E | 7283 hCoV-19/Germany/NW-MPP-8/2020\|EPI_ISL_487430\|2020- | .................... | .................... | .................... | .................... | .................... | .................... |
| E | 7284 hCoV-19/Germany/NW-MPP-9/2020\|EPI_ISL_487431\|2020- | .................... | .................... | .................... | .................... | .................... | .................... |
| E | 7285 hCoV-19/Belgium/ITM_C2184_2/2020\|EPI_ISL_487432\|20 | .................... | .................... | .................... | .................... | .................... | .................... |
| E | 7286 hCoV-19/Belgium/ITM_C2181/2020\|EPI_ISL_487433\|2020 | .................... | .................... | .................... | .................... | .................... | .................... |
| E | 7287 hCoV-19/Belgium/ITM_C2182/2020\|EPI_ISL_487434\|2020 | .................... | .................... | .................... | .................... | .................... | .................... |
| E | 7288 hCoV-19/Belgium/ITM_C2183/2020\|EPI_ISL_487435\|2020 | .................... | .................... | .................... | .................... | .................... | .................... |
| E | 7289 hCoV-19/Belgium/ITM_C2185/2020\|EPI_ISL_487436\|2020 | .................... | .................... | .................... | .................... | .................... | .................... |
| E | 7290 hCoV-19/Mali/M002593/2020\|EPI_ISL_487446\|2020-04-0 | .................... | .................... | .................... | .................... | .................... | .................... |
| E | 7291 hCoV-19/Mali/M002615/2020\|EPI_ISL_487447\|2020-04-0 | .................... | .................... | .................... | .................... | .................... | .................... |
| E | 7292 hCoV-19/Mali/M002616/2020\|EPI_ISL_487448\|2020-04-0 | .................... | .................... | .................... | .................... | .................... | .................... |
| E | 7293 hCoV-19/Mali/M002618/2020\|EPI_ISL_487449\|2020-04-0 | .................... | .................... | .................... | .................... | .................... | .................... |
| E | 7294 hCoV-19/Mali/M002644/2020\|EPI_ISL_487450\|2020-04-0 | .................... | .................... | .................... | .................... | .................... | .................... |
| E | 7295 hCoV-19/Mali/M002659/2020\|EPI_ISL_487451\|2020-04-0 | .................... | .................... | .................... | .................... | .................... | .................... |
| E | 7296 hCoV-19/Mali/M002663/2020\|EPI_ISL_487452\|2020-04-0 | .................... | .................... | .................... | .................... | .................... | .................... |
| E | 7297 hCoV-19/Mali/M002667/2020\|EPI_ISL_487453\|2020-04-1 | .................... | .................... | .................... | .................... | .................... | .................... |
| E | 7298 hCoV-19/Mali/M002672/2020\|EPI_ISL_487454\|2020-04-1 | .................... | .................... | .................... | .................... | .................... | .................... |
| E | 7299 hCoV-19/Mali/M002673/2020\|EPI_ISL_487455\|2020-04-1 | .................... | .................... | .................... | .................... | .................... | .................... |
| E | 7300 hCoV-19/Mali/M002675/2020\|EPI_ISL_487456\|2020-04-1 | .................... | .................... | .................... | .................... | .................... | .................... |
| E | 7301 hCoV-19/Mali/M002698/2020\|EPI_ISL_487457\|2020-04-1 | .................... | .................... | .................... | .................... | .................... | .................... |
| E | 7302 hCoV-19/Mali/M002700/2020\|EPI_ISL_487458\|2020-04-1 | .................... | .................... | .................... | .................... | .................... | .................... |
| E | 7303 hCoV-19/Mali/M002703/2020\|EPI_ISL_487459\|2020-04-1 | .................... | .................... | .................... | .................... | .................... | .................... |
| E | 7304 hCoV-19/Mali/M002704/2020\|EPI_ISL_487460\|2020-04-1 | .................... | .................... | .................... | .................... | .................... | .................... |
| E | 7305 hCoV-19/Mali/M002707/2020\|EPI_ISL_487461\|2020-04-1 | .................... | .................... | .................... | .................... | .................... | .................... |
| E | 7306 hCoV-19/Mali/M002758/2020\|EPI_ISL_487462\|2020-04-1 | .................... | .................... | .................... | .................... | .................... | .................... |
| E | 7307 hCoV-19/Mali/M002823/2020\|EPI_ISL_487463\|2020-04-1 | .................... | .................... | .................... | .................... | .................... | .................... |
| E | 7308 hCoV-19/Mali/M002824/2020\|EPI_ISL_487464\|2020-04-1 | .................... | .................... | .................... | .................... | .................... | .................... |
| E | 7309 hCoV-19/Mali/M002826/2020\|EPI_ISL_487465\|2020-04-1 | .................... | .................... | .................... | .................... | .................... | .................... |
| E | 7310 hCoV-19/Mali/M002830/2020\|EPI_ISL_487466\|2020-04-1 | .................... | .................... | .................... | .................... | .................... | .................... |
| E | 7311 hCoV-19/England/NORT-2834CB/2020\|EPI_ISL_488700\|20 | .................... | .................... | .................... | .................... | .................... | .................... |
| E | 7312 hCoV-19/England/NORT-2849AD/2020\|EPI_ISL_488701\|20 | .................... | .................... | .................... | .................... | .................... | .................... |

| | | | | | | | | |
|---|---|---|---|---|---|---|---|---|
| E | 7313 | hCoV-19/England/NORT-295156/2020\|EPI_ISL_488702\|20 | .................... | .................... | ..................... | .................... | .................... | .................... |
| E | 7314 | hCoV-19/England/NORT-2957DC/2020\|EPI_ISL_488703\|20 | .................... | .................... | ..................... | .................... | .................... | .................... |
| E | 7315 | hCoV-19/England/NORT-295ED7/2020\|EPI_ISL_488704\|20 | .................... | .................... | ..................... | .................... | .................... | .................... |
| E | 7316 | hCoV-19/England/NORT-283917/2020\|EPI_ISL_488705\|20 | .................... | .................... | ..................... | .................... | .................... | .................... |
| E | 7317 | hCoV-19/England/NORT-295192/2020\|EPI_ISL_488706\|20 | .................... | .................... | ..................... | .................... | .................... | .................... |
| E | 7318 | hCoV-19/England/NORT-28421B/2020\|EPI_ISL_488707\|20 | .................... | .................... | ..................... | .................... | .................... | .................... |
| E | 7319 | hCoV-19/England/NORT-2839AE/2020\|EPI_ISL_488708\|20 | .................... | .................... | ..................... | .................... | .................... | .................... |
| E | 7320 | hCoV-19/England/NORT-29561B/2020\|EPI_ISL_488709\|20 | .................... | .................... | ..................... | .................... | .................... | .................... |
| E | 7321 | hCoV-19/England/NORT-295B67/2020\|EPI_ISL_488710\|20 | .................... | ....A............... | ..................... | .................... | .................... | .................... |
| E | 7322 | hCoV-19/England/NORT-29594F/2020\|EPI_ISL_488711\|20 | .................... | .................... | ..................... | .................... | .................... | .................... |
| E | 7323 | hCoV-19/England/NORT-2953AB/2020\|EPI_ISL_488712\|20 | .................... | .................... | ..................... | .................... | .................... | .................... |
| E | 7324 | hCoV-19/England/NORT-2831F1/2020\|EPI_ISL_488713\|20 | .................... | .................... | ..................... | .................... | .................... | .................... |
| E | 7325 | hCoV-19/England/NORT-28454F/2020\|EPI_ISL_488714\|20 | .................... | .................... | ..................... | .................... | .................... | .................... |
| E | 7326 | hCoV-19/England/NORT-284C1D/2020\|EPI_ISL_488715\|20 | .................... | .................... | ..................... | .................... | .................... | .................... |
| E | 7327 | hCoV-19/England/NORT-28257E/2020\|EPI_ISL_488716\|20 | .................... | .................... | ..................... | .................... | .................... | .................... |
| E | 7328 | hCoV-19/England/NORT-294DAE/2020\|EPI_ISL_488717\|20 | .................... | .................... | ..................... | .................... | .................... | .................... |
| E | 7329 | hCoV-19/England/NORT-2827E1/2020\|EPI_ISL_488718\|20 | .................... | .................... | ..................... | .................... | .................... | .................... |
| E | 7330 | hCoV-19/England/NORT-294E41/2020\|EPI_ISL_488719\|20 | .................... | .................... | ..................... | .................... | .................... | .................... |
| E | 7331 | hCoV-19/England/NORT-2833BF/2020\|EPI_ISL_488720\|20 | .................... | .................... | ..................... | .................... | .................... | .................... |
| E | 7332 | hCoV-19/England/NORT-29599A/2020\|EPI_ISL_488721\|20 | .................... | .................... | ..................... | .................... | .................... | .................... |
| E | 7333 | hCoV-19/England/NORT-295D25/2020\|EPI_ISL_488722\|20 | .................... | .................... | ..................... | .................... | .................... | .................... |
| E | 7334 | hCoV-19/England/NORT-295DCB/2020\|EPI_ISL_488723\|20 | .................... | .................... | ..................... | .................... | .................... | .................... |
| E | 7335 | hCoV-19/England/NORT-282B9A/2020\|EPI_ISL_488724\|20 | .................... | .................... | ..................... | .................... | .................... | .................... |
| E | 7336 | hCoV-19/England/NORT-284846/2020\|EPI_ISL_488725\|20 | .................... | .................... | ..................... | .................... | .................... | .................... |
| E | 7337 | hCoV-19/England/NORT-282AD9/2020\|EPI_ISL_488726\|20 | .................... | .................... | ..................... | .................... | .................... | .................... |
| E | 7338 | hCoV-19/England/NORT-294F99/2020\|EPI_ISL_488727\|20 | .................... | .................... | ..................... | .................... | .................... | .................... |
| E | 7339 | hCoV-19/England/NORT-2827D2/2020\|EPI_ISL_488728\|20 | .................... | .................... | ..................... | .................... | .................... | .................... |
| E | 7340 | hCoV-19/England/NORT-2844BB/2020\|EPI_ISL_488729\|20 | .................... | .................... | ..................... | .................... | .................... | .................... |
| E | 7341 | hCoV-19/England/NORT-282FCB/2020\|EPI_ISL_488730\|20 | .................... | .................... | ..................... | .................... | .................... | .................... |
| E | 7342 | hCoV-19/England/NORT-28207D/2020\|EPI_ISL_488731\|20 | .................... | .................... | ..................... | .................... | .................... | .................... |
| E | 7343 | hCoV-19/England/NORT-2950D1/2020\|EPI_ISL_488732\|20 | .................... | .................... | ..................... | .................... | .................... | .................... |
| E | 7344 | hCoV-19/England/NORT-294BE0/2020\|EPI_ISL_488733\|20 | .................... | .................... | ..................... | .................... | .................... | .................... |
| E | 7345 | hCoV-19/England/NORT-28348F/2020\|EPI_ISL_488734\|20 | .................... | .................... | ..................... | .................... | .................... | .................... |
| E | 7346 | hCoV-19/England/NORT-283856/2020\|EPI_ISL_488735\|20 | .................... | .................... | ..................... | .................... | .................... | .................... |
| E | 7347 | hCoV-19/England/NORT-295745/2020\|EPI_ISL_488736\|20 | .................... | .................... | ..................... | .................... | .................... | .................... |
| E | 7348 | hCoV-19/England/NORT-28414B/2020\|EPI_ISL_488737\|20 | .................... | .................... | ..................... | .................... | .................... | .................... |
| E | 7349 | hCoV-19/England/NORT-295E8C/2020\|EPI_ISL_488738\|20 | .................... | .................... | ..................... | .................... | .................... | .................... |
| E | 7350 | hCoV-19/England/NORT-28213E/2020\|EPI_ISL_488739\|20 | .................... | .................... | ..................... | .................... | .................... | .................... |
| E | 7351 | hCoV-19/England/NORT-282F34/2020\|EPI_ISL_488740\|20 | .................... | .................... | ..................... | .................... | .................... | .................... |
| E | 7352 | hCoV-19/England/NORT-28314C/2020\|EPI_ISL_488741\|20 | .................... | .................... | ..................... | .................... | .................... | .................... |
| E | 7353 | hCoV-19/England/NORT-283D57/2020\|EPI_ISL_488742\|20 | .................... | .................... | ..................... | .................... | .................... | .................... |
| E | 7354 | hCoV-19/England/NORT-284451/2020\|EPI_ISL_488743\|20 | .................... | .................... | ..................... | .................... | .................... | .................... |
| E | 7355 | hCoV-19/England/NORT-2842FD/2020\|EPI_ISL_488744\|20 | .................... | .................... | ..................... | .................... | .................... | .................... |
| E | 7356 | hCoV-19/England/NORT-283B5D/2020\|EPI_ISL_488745\|20 | .................... | .................... | ..................... | .................... | .................... | .................... |
| E | 7357 | hCoV-19/England/NORT-294FA8/2020\|EPI_ISL_488746\|20 | .................... | .................... | ..................... | .................... | .................... | .................... |
| E | 7358 | hCoV-19/England/NORT-281991/2020\|EPI_ISL_488747\|20 | .................... | .................... | ..................... | .................... | .................... | .................... |
| E | 7359 | hCoV-19/England/NORT-28125A/2020\|EPI_ISL_488748\|20 | .................... | .................... | ..................... | .................... | .................... | .................... |
| E | 7360 | hCoV-19/England/NORT-281506/2020\|EPI_ISL_488749\|20 | .................... | .................... | ..................... | .................... | .................... | .................... |
| E | 7361 | hCoV-19/England/NORT-2814EB/2020\|EPI_ISL_488750\|20 | .................... | .................... | ..................... | .................... | .................... | .................... |
| E | 7362 | hCoV-19/England/NORT-283B20/2020\|EPI_ISL_488751\|20 | .................... | .................... | ..................... | .................... | .................... | .................... |
| E | 7363 | hCoV-19/England/NORT-2818EF/2020\|EPI_ISL_488752\|20 | .................... | .................... | ..................... | .................... | .................... | .................... |
| E | 7364 | hCoV-19/England/NORT-281C7A/2020\|EPI_ISL_488753\|20 | .................... | .................... | ..................... | .................... | .................... | .................... |
| E | 7365 | hCoV-19/England/NORT-281F53/2020\|EPI_ISL_488754\|20 | .................... | .................... | ..................... | .................... | .................... | .................... |
| E | 7366 | hCoV-19/England/NORT-281876/2020\|EPI_ISL_488755\|20 | .................... | .................... | ..................... | .................... | .................... | .................... |
| E | 7367 | hCoV-19/England/NORT-2812A5/2020\|EPI_ISL_488756\|20 | .................... | .................... | ..................... | .................... | .................... | .................... |
| E | 7368 | hCoV-19/England/NORT-2816C7/2020\|EPI_ISL_488757\|20 | .................... | .................... | ..................... | .................... | .................... | .................... |
| E | 7369 | hCoV-19/England/NORT-2813B1/2020\|EPI_ISL_488758\|20 | .................... | .................... | ..................... | .................... | .................... | .................... |
| E | 7370 | hCoV-19/England/NORT-281A8F/2020\|EPI_ISL_488759\|20 | .................... | .................... | ..................... | .................... | .................... | .................... |
| E | 7371 | hCoV-19/England/NORT-281023/2020\|EPI_ISL_488760\|20 | .................... | .................... | ..................... | .................... | .................... | .................... |
| E | 7372 | hCoV-19/England/NORT-283DA2/2020\|EPI_ISL_488761\|20 | .................... | .................... | ..................... | .................... | .................... | .................... |
| E | 7373 | hCoV-19/England/NORT-28166D/2020\|EPI_ISL_488762\|20 | .................... | .................... | ..................... | .................... | .................... | .................... |
| E | 7374 | hCoV-19/England/NORT-281700/2020\|EPI_ISL_488763\|20 | .................... | .................... | ..................... | .................... | .................... | .................... |
| E | 7375 | hCoV-19/England/NORT-281357/2020\|EPI_ISL_488764\|20 | .................... | .................... | ..................... | .................... | .................... | .................... |
| E | 7376 | hCoV-19/England/NORT-2816E5/2020\|EPI_ISL_488765\|20 | .................... | .................... | ..................... | .................... | .................... | .................... |
| E | 7377 | hCoV-19/England/NORT-28157F/2020\|EPI_ISL_488766\|20 | .................... | .................... | ..................... | .................... | .................... | .................... |
| E | 7378 | hCoV-19/England/NORT-2814AF/2020\|EPI_ISL_488767\|20 | .................... | .................... | ..................... | .................... | .................... | .................... |
| E | 7379 | hCoV-19/England/NORT-281DFF/2020\|EPI_ISL_488768\|20 | .................... | .................... | ..................... | .................... | .................... | .................... |
| E | 7380 | hCoV-19/England/NORT-281858/2020\|EPI_ISL_488769\|20 | .................... | .................... | ..................... | .................... | .................... | .................... |
| E | 7381 | hCoV-19/England/NORT-281348/2020\|EPI_ISL_488771\|20 | .................... | .................... | ..................... | .................... | .................... | .................... |
| E | 7382 | hCoV-19/England/NORT-281445/2020\|EPI_ISL_488772\|20 | .................... | .................... | ..................... | .................... | .................... | .................... |
| E | 7383 | hCoV-19/England/NORT-281919/2020\|EPI_ISL_488773\|20 | .................... | .................... | ..................... | .................... | .................... | .................... |
| E | 7384 | hCoV-19/England/NORT-281621/2020\|EPI_ISL_488774\|20 | .................... | .................... | ..................... | .................... | .................... | .................... |
| E | 7385 | hCoV-19/England/NORT-281481/2020\|EPI_ISL_488775\|20 | .................... | .................... | ..................... | .................... | .................... | .................... |
| E | 7386 | hCoV-19/England/NORT-28404E/2020\|EPI_ISL_488776\|20 | .................... | .................... | ..................... | .................... | .................... | .................... |
| E | 7387 | hCoV-19/England/NORT-281937/2020\|EPI_ISL_488777\|20 | .................... | .................... | ..................... | .................... | .................... | .................... |
| E | 7388 | hCoV-19/England/NORT-2819BF/2020\|EPI_ISL_488778\|20 | .................... | .................... | ..................... | .................... | .................... | .................... |
| E | 7389 | hCoV-19/England/NORT-2814FA/2020\|EPI_ISL_488779\|20 | .................... | .................... | ..................... | .................... | .................... | .................... |
| E | 7390 | hCoV-19/England/NORT-281F08/2020\|EPI_ISL_488780\|20 | .................... | .................... | ..................... | .................... | .................... | .................... |
| E | 7391 | hCoV-19/England/NORT-281F35/2020\|EPI_ISL_488782\|20 | .................... | .................... | ..................... | .................... | .................... | .................... |
| E | 7392 | hCoV-19/England/NORT-2818A3/2020\|EPI_ISL_488783\|20 | .................... | .................... | ..................... | .................... | .................... | .................... |

| | | | | | | | |
|---|---|---|---|---|---|---|---|
| E | 7393 hCoV-19/England/NORT-283E72/2020\|EPI_ISL_488784\|20 | ···················· | ····T··············· | ···················· | ···················· | ···················· | ···················· |
| E | 7394 hCoV-19/England/NORT-2814DC/2020\|EPI_ISL_488785\|20 | ···················· | ···················· | ···················· | ···················· | ···················· | ···················· |
| E | 7395 hCoV-19/England/NORT-281454/2020\|EPI_ISL_488786\|20 | ···················· | ···················· | ···················· | ···················· | ···················· | ···················· |
| E | 7396 hCoV-19/England/NORT-281955/2020\|EPI_ISL_488787\|20 | ···················· | ···················· | ···················· | ···················· | ···················· | ···················· |
| E | 7397 hCoV-19/England/NORT-281050/2020\|EPI_ISL_488788\|20 | ···················· | ···················· | ···················· | ···················· | ···················· | ···················· |
| E | 7398 hCoV-19/England/NORT-28171F/2020\|EPI_ISL_488789\|20 | ···················· | ···················· | ···················· | ···················· | ···················· | ···················· |
| E | 7399 hCoV-19/England/NORT-281BE6/2020\|EPI_ISL_488790\|20 | ···················· | ···················· | ···················· | ···················· | ···················· | ···················· |
| E | 7400 hCoV-19/England/NORT-2841D2/2020\|EPI_ISL_488791\|20 | ···················· | ···················· | ···················· | ···················· | ···················· | ···················· |
| E | 7401 hCoV-19/England/NORT-281612/2020\|EPI_ISL_488792\|20 | ···················· | ···················· | ···················· | ···················· | ···················· | ···················· |
| E | 7402 hCoV-19/England/NORT-28183A/2020\|EPI_ISL_488793\|20 | ···················· | ···················· | ···················· | ···················· | ···················· | ···················· |
| E | 7403 hCoV-19/England/NORT-28172E/2020\|EPI_ISL_488794\|20 | ···················· | ···················· | ···················· | ···················· | ···················· | ···················· |
| E | 7404 hCoV-19/England/NORT-281E1A/2020\|EPI_ISL_488795\|20 | ···················· | ···················· | ···················· | ···················· | ···················· | ···················· |
| E | 7405 hCoV-19/England/NORT-281D59/2020\|EPI_ISL_488796\|20 | ···················· | ···················· | ···················· | ···················· | ···················· | ···················· |
| E | 7406 hCoV-19/England/NORT-281463/2020\|EPI_ISL_488797\|20 | ···················· | ···················· | ···················· | ···················· | ···················· | ···················· |
| E | 7407 hCoV-19/England/NORT-2817A6/2020\|EPI_ISL_488798\|20 | ···················· | ···················· | ···················· | ···················· | ···················· | ···················· |
| E | 7408 hCoV-19/England/NORT-28173D/2020\|EPI_ISL_488799\|20 | ···················· | ···················· | ···················· | ···················· | ···················· | ···················· |
| E | 7409 hCoV-19/USA/CA-ALSR-1382/2020\|EPI_ISL_494573\|2020- | ···················· | ···················· | ···················· | ···················· | ···················· | ···················· |
| E | 7410 hCoV-19/USA/CA-ALSR-1384/2020\|EPI_ISL_494575\|2020- | ···················· | ···················· | ···················· | ···················· | ···················· | ···················· |
| E | 7411 hCoV-19/USA/CA-ALSR-1387/2020\|EPI_ISL_494577\|2020- | ···················· | ·A·················· | ···················· | ···················· | ···················· | ···················· |
| E | 7412 hCoV-19/USA/CA-ALSR-1388/2020\|EPI_ISL_494578\|2020- | ···················· | ·A·················· | ···················· | ···················· | ···················· | ···················· |
| E | 7413 hCoV-19/USA/CA-ALSR-1390/2020\|EPI_ISL_494579\|2020- | ···················· | ·A·················· | ···················· | ···················· | ···················· | ···················· |
| E | 7414 hCoV-19/USA/CA-ALSR-1398/2020\|EPI_ISL_494584\|2020- | ···················· | ·A·················· | ···················· | ···················· | ···················· | ···················· |
| E | 7415 hCoV-19/USA/CA-ALSR-1407/2020\|EPI_ISL_494585\|2020- | ···················· | ·A·················· | ···················· | ···················· | ···················· | ···················· |
| E | 7416 hCoV-19/USA/CA-ALSR-1410/2020\|EPI_ISL_494586\|2020- | ···················· | ·A·················· | ···················· | ···················· | ···················· | ···················· |
| E | 7417 hCoV-19/USA/CA-ALSR-1411/2020\|EPI_ISL_494587\|2020- | ···················· | ·A·················· | ···················· | ···················· | ···················· | ···················· |
| E | 7418 hCoV-19/USA/CA-ALSR-1413/2020\|EPI_ISL_494588\|2020- | ···················· | ·A·················· | ···················· | ···················· | ···················· | ···················· |
| E | 7419 hCoV-19/USA/CA-ALSR-1415/2020\|EPI_ISL_494589\|2020- | ···················· | ·A·················· | ···················· | ···················· | ···················· | ···················· |
| E | 7420 hCoV-19/USA/CA-ALSR-1416/2020\|EPI_ISL_494590\|2020- | ···················· | ·A·················· | ···················· | ···················· | ···················· | ···················· |
| E | 7421 hCoV-19/USA/CA-ALSR-1418/2020\|EPI_ISL_494591\|2020- | ···················· | ·A·················· | ···················· | ···················· | ···················· | ···················· |
| E | 7422 hCoV-19/USA/CA-ALSR-1419/2020\|EPI_ISL_494592\|2020- | ···················· | ···················· | ···················· | ···················· | ···················· | ···················· |
| E | 7423 hCoV-19/USA/CA-ALSR-1422/2020\|EPI_ISL_494593\|2020- | ···················· | ·A·················· | ···················· | ···················· | ···················· | ···················· |
| E | 7424 hCoV-19/USA/CA-ALSR-1425/2020\|EPI_ISL_494594\|2020- | ···················· | ·A·················· | ···················· | ···················· | ···················· | ···················· |
| E | 7425 hCoV-19/India/OR-ILSCV35476/2020\|EPI_ISL_481200\|20 | ···················· | ···················· | ···················· | ···················· | ···················· | ···················· |
| E | 7426 hCoV-19/India/OR-ILSCV35482/2020\|EPI_ISL_481201\|20 | ···················· | ···················· | ···················· | ···················· | ···················· | ···················· |
| E | 7427 hCoV-19/India/OR-ILSCV35574/2020\|EPI_ISL_481202\|20 | ···················· | ···················· | ···················· | ···················· | ···················· | ···················· |
| E | 7428 hCoV-19/India/OR-ILSCV35614/2020\|EPI_ISL_481203\|20 | ···················· | ···················· | ···················· | ···················· | ······T············· | ···················· |
| E | 7429 hCoV-19/India/OR-ILSCV35615/2020\|EPI_ISL_481204\|20 | ···················· | ···················· | ···················· | ···················· | ······T············· | ···················· |
| E | 7430 hCoV-19/India/OR-ILSCV35750/2020\|EPI_ISL_481205\|20 | ···················· | ···················· | ···················· | ···················· | ···················· | ···················· |
| E | 7431 hCoV-19/Belgium/UGent-124/2020\|EPI_ISL_481221\|2020 | ···················· | ···················· | ···················· | ···················· | ···················· | ···················· |
| E | 7434 hCoV-19/Belgium/UGent-129/2020\|EPI_ISL_481225\|2020 | ···················· | ···················· | ···················· | ···················· | ···················· | ···················· |
| E | 7435 hCoV-19/Belgium/UGent-133/2020\|EPI_ISL_481227\|2020 | ···················· | ···················· | ···················· | ···················· | ···················· | ···················· |
| E | 7438 hCoV-19/Belgium/UGent-140/2020\|EPI_ISL_481231\|2020 | ···················· | ···················· | ···················· | ···················· | ···················· | ···················· |
| E | 7440 hCoV-19/Belgium/UGent-142/2020\|EPI_ISL_481233\|2020 | ···················· | ···················· | ···················· | ···················· | ···················· | ···················· |
| E | 7441 hCoV-19/Senegal/3331/2020\|EPI_ISL_481235\|2020-04-0 | ···················· | ···················· | ···················· | ···················· | ···················· | ···················· |
| E | 7442 hCoV-19/Senegal/5718/2020\|EPI_ISL_481236\|2020-04-1 | ···················· | ···················· | ···················· | ···················· | ···················· | ···················· |
| E | 7443 hCoV-19/Senegal/5766/2020\|EPI_ISL_481237\|2020-04-1 | ···················· | ···················· | ···················· | ···················· | ···················· | ···················· |
| E | 7444 hCoV-19/Senegal/5785/2020\|EPI_ISL_481239\|2020-04-1 | ···················· | ···················· | ···················· | ···················· | ···················· | ···················· |
| E | 7445 hCoV-19/Senegal/5971/2020\|EPI_ISL_481240\|2020-04-1 | ···················· | ···················· | ···················· | ···················· | ···················· | ···················· |
| E | 7446 hCoV-19/USA/MN-MDH-1200/2020\|EPI_ISL_481241\|2020-0 | ···················· | ·A·················· | ···················· | ···················· | ···················· | ···················· |
| E | 7447 hCoV-19/USA/MN-MDH-1224/2020\|EPI_ISL_481242\|2020-0 | ···················· | ···················· | ···················· | ···················· | ···················· | ···················· |
| E | 7448 hCoV-19/Senegal/5986/2020\|EPI_ISL_481243\|2020-04-1 | ···················· | ···················· | ···················· | ···················· | ···················· | ···················· |
| E | 7449 hCoV-19/Ecuador/USFQ-026/2020\|EPI_ISL_481244\|2020- | ···················· | ···················· | ···················· | ···················· | ···················· | ···················· |
| E | 7450 hCoV-19/Ecuador/USFQ-039/2020\|EPI_ISL_481245\|2020- | ···················· | ···················· | ···················· | ···················· | ···················· | ···················· |
| E | 7451 hCoV-19/Ecuador/USFQ-054/2020\|EPI_ISL_481246\|2020- | ···················· | ···················· | ···················· | ···················· | ···················· | ···················· |
| E | 7452 hCoV-19/Ecuador/USFQ-065/2020\|EPI_ISL_481247\|2020- | ···················· | ···················· | ···················· | ···················· | ···················· | ···················· |
| E | 7453 hCoV-19/Ecuador/USFQ-066/2020\|EPI_ISL_481248\|2020- | ···················· | ···················· | ···················· | ···················· | ···················· | ···················· |
| E | 7454 hCoV-19/Japan/NGS-IA-1/2020\|EPI_ISL_481251\|2020-03 | ···················· | ···················· | ···················· | ···················· | ···················· | ···················· |
| E | 7455 hCoV-19/Germany/BW-RKI-N-0001/2020\|EPI_ISL_481253\| | ···················· | ···················· | ···················· | ···················· | ···················· | ···················· |
| E | 7456 hCoV-19/Japan/NGS-SC-1/2020\|EPI_ISL_481254\|2020-04 | ···················· | ···················· | ···················· | ···················· | ···················· | ···················· |
| E | 7457 hCoV-19/Japan/NGS-CD-010/2020\|EPI_ISL_481255\|2020- | ···················· | ···················· | ···················· | ···················· | ···················· | ···················· |
| E | 7458 hCoV-19/Germany/NW-RKI-N-0002/2020\|EPI_ISL_481256\| | ···················· | ···················· | ···················· | ···················· | ···················· | ···················· |
| E | 7459 hCoV-19/Japan/NGS-CD-024/2020\|EPI_ISL_481257\|2020- | ···················· | ···················· | ···················· | ···················· | ···················· | ···················· |
| E | 7460 hCoV-19/Japan/NGS-CD-107/2020\|EPI_ISL_481258\|2020- | ···················· | ···················· | ···················· | ···················· | ···················· | ···················· |
| E | 7461 hCoV-19/Japan/NGS-CD-235/2020\|EPI_ISL_481259\|2020- | ···················· | ···················· | ···················· | ···················· | ···················· | ···················· |
| E | 7462 hCoV-19/Japan/NGS-CD-292/2020\|EPI_ISL_481260\|2020- | ···················· | ···················· | ···················· | ···················· | ···················· | ···················· |
| E | 7463 hCoV-19/Japan/NGS-CD-361/2020\|EPI_ISL_481261\|2020- | ···················· | ···················· | ···················· | ···················· | ···················· | ···················· |
| E | 7464 hCoV-19/Germany/BY-RKI-N-0003/2020\|EPI_ISL_481262\| | ···················· | ···················· | ···················· | ···················· | ···················· | ···················· |
| E | 7465 hCoV-19/USA/MD-MDH-0030/2020\|EPI_ISL_481275\|2020-0 | ···················· | ···················· | ···················· | ···················· | ···················· | ···················· |
| E | 7466 hCoV-19/USA/MD-MDH-0033/2020\|EPI_ISL_481278\|2020-0 | ···················· | ···················· | ···················· | ···················· | ···················· | ···················· |
| E | 7467 hCoV-19/USA/MD-MDH-0034/2020\|EPI_ISL_481279\|2020-0 | ···················· | ···················· | ···················· | ···················· | ···················· | ···················· |
| E | 7468 hCoV-19/India/DL-NCDC1892-CSIR-IGIB/2020\|EPI_ISL_4 | ···················· | ···················· | ···················· | ···················· | ···················· | ···················· |
| E | 7469 hCoV-19/India/DL-NCDC1895-CSIR-IGIB/2020\|EPI_ISL_4 | ···················· | ·A·················· | ···················· | ···················· | ···················· | ···A················ |
| E | 7470 hCoV-19/India/DL-NCDC1934-CSIR-IGIB/2020\|EPI_ISL_4 | ···················· | ·R·················· | ···················· | ···················· | ···················· | ···N················ |
| E | 7471 hCoV-19/India/DL-NCDC1957-CSIR-IGIB/2020\|EPI_ISL_4 | ···················· | ···················· | ···················· | ···················· | ···················· | ···················· |
| E | 7472 hCoV-19/India/DL-NCDC2037-CSIR-IGIB/2020\|EPI_ISL_4 | ···················· | ···················· | ···················· | ···················· | ···················· | ···················· |
| E | 7473 hCoV-19/India/DL-NCDC2058-CSIR-IGIB/2020\|EPI_ISL_4 | ···················· | ···················· | ···················· | ···················· | ···················· | ···················· |
| E | 7474 hCoV-19/India/DL-NCDC2214-CSIR-IGIB/2020\|EPI_ISL_4 | ···················· | ···················· | ···················· | ···················· | ···················· | ···················· |
| E | 7475 hCoV-19/India/DL-NCDC2283-CSIR-IGIB/2020\|EPI_ISL_4 | ···················· | ·A·················· | ···················· | ···················· | ···················· | ···A················ |
| E | 7476 hCoV-19/India/DL-NCDC2293-CSIR-IGIB/2020\|EPI_ISL_4 | ···················· | ·N·················· | ···················· | ···················· | ···················· | ···N················ |
| E | 7477 hCoV-19/India/DL-NCDC2334-CSIR-IGIB/2020\|EPI_ISL_4 | ···················· | ···················· | ···················· | ···················· | ···················· | ···················· |

| | | | | | | | | |
|---|---|---|---|---|---|---|---|---|
| E | 7478 | hCoV-19/India/DL-NCDC3652-CSIR-IGIB/2020\|EPI_ISL_4 | ···················· | ·A·················· | ····················· | ···················· | ···················· | ···A················ |
| E | 7479 | hCoV-19/India/DL-NCDC3820-CSIR-IGIB/2020\|EPI_ISL_4 | ···················· | ·A·················· | ····················· | ···················· | ···················· | ···N················ |
| E | 7480 | hCoV-19/Hangzhou/HZCDC6111/2020\|EPI_ISL_482575\|202 | ···················· | ···················· | ····················· | ···················· | ···················· | ···················· |
| E | 7481 | hCoV-19/Hangzhou/HZCDC6224/2020\|EPI_ISL_482576\|202 | ···················· | ···················· | ····················· | ···················· | ···················· | ···················· |
| E | 7482 | hCoV-19/Hangzhou/HZCDC6228/2020\|EPI_ISL_482577\|202 | ···················· | ···················· | ····················· | ···················· | ···················· | ···················· |
| E | 7483 | hCoV-19/Hangzhou/HZCDC6445/2020\|EPI_ISL_482578\|202 | ···················· | ···················· | ····················· | ···················· | ···················· | ···················· |
| E | 7484 | hCoV-19/Hangzhou/HZCDC6948/2020\|EPI_ISL_482579\|202 | ···················· | ···················· | ····················· | ···················· | ···················· | ···················· |
| E | 7485 | hCoV-19/Hangzhou/HZCDC7019/2020\|EPI_ISL_482580\|202 | ···················· | ···················· | ····················· | ···················· | ···················· | ···················· |
| E | 7486 | hCoV-19/Hangzhou/HZCDC7155/2020\|EPI_ISL_482581\|202 | ···················· | ···················· | ····················· | ···················· | ···················· | ···················· |
| E | 7487 | hCoV-19/Hangzhou/HZCDCp145/2020\|EPI_ISL_482582\|202 | ···················· | ···················· | ····················· | ···················· | ···················· | ···················· |
| E | 7488 | hCoV-19/Hangzhou/HZCDC7184F/2020\|EPI_ISL_482583\|20 | ···················· | ···················· | ····················· | ···················· | ···················· | ···················· |
| E | 7489 | hCoV-19/Hangzhou/HZCDC7328/2020\|EPI_ISL_482584\|202 | ···················· | ···················· | ····················· | ···················· | ···················· | ···················· |
| E | 7490 | hCoV-19/Hangzhou/HZCDC7363/2020\|EPI_ISL_482585\|202 | ···················· | ···················· | ····················· | ···················· | ···················· | ···················· |
| E | 7491 | hCoV-19/Hangzhou/HZCDC7378/2020\|EPI_ISL_482586\|202 | ···················· | ···················· | ····················· | ···················· | ···················· | ···················· |
| E | 7492 | hCoV-19/India/DL-NCDC3832-CSIR-IGIB/2020\|EPI_ISL_4 | ···················· | ·A·················· | ····················· | ···················· | ···················· | ···A················ |
| E | 7493 | hCoV-19/India/DL-NCDC4402-CSIR-IGIB/2020\|EPI_ISL_4 | ···················· | ·A·················· | ····················· | ···················· | ···················· | ···A················ |
| E | 7494 | hCoV-19/India/DL-NCDC4462-CSIR-IGIB/2020\|EPI_ISL_4 | ···················· | ·A·················· | ····················· | ···················· | ···················· | ···A················ |
| E | 7495 | hCoV-19/India/GJ-GBRC254a/2020\|EPI_ISL_483840\|2020 | ···················· | ·A·················· | ····················· | ···················· | ···················· | ···A················ |
| E | 7496 | hCoV-19/India/GJ-GBRC254b/2020\|EPI_ISL_483841\|2020 | ···················· | ·A·················· | ····················· | ···················· | ···················· | ···A················ |
| E | 7497 | hCoV-19/India/GJ-GBRC255a/2020\|EPI_ISL_483842\|2020 | ···················· | ·A·················· | ····················· | ···················· | ···················· | ···A················ |
| E | 7498 | hCoV-19/India/GJ-GBRC255b/2020\|EPI_ISL_483843\|2020 | ···················· | ·A·················· | ····················· | ···················· | ···················· | ···A················ |
| E | 7499 | hCoV-19/India/GJ-GBRC256a/2020\|EPI_ISL_483844\|2020 | ···················· | ···················· | ····················· | ···················· | ···················· | ···················· |
| E | 7500 | hCoV-19/India/GJ-GBRC256b/2020\|EPI_ISL_483845\|2020 | ···················· | ···················· | ····················· | ···················· | ···················· | ···A················ |
| E | 7501 | hCoV-19/India/GJ-GBRC257a/2020\|EPI_ISL_483846\|2020 | ···················· | ·A·················· | ····················· | ···················· | ···················· | ···A················ |
| E | 7502 | hCoV-19/India/GJ-GBRC257b/2020\|EPI_ISL_483847\|2020 | ···················· | ·A·················· | ····················· | ···················· | ···················· | ···A················ |
| E | 7503 | hCoV-19/India/GJ-GBRC258a/2020\|EPI_ISL_483848\|2020 | ···················· | ···················· | ····················· | ···················· | ···················· | ···A················ |
| E | 7504 | hCoV-19/India/GJ-GBRC258b/2020\|EPI_ISL_483849\|2020 | ···················· | ·A·················· | ····················· | ···················· | ···················· | ···················· |
| E | 7505 | hCoV-19/India/GJ-GBRC259/2020\|EPI_ISL_483850\|2020- | ···················· | ···················· | ····················· | ···················· | ···················· | ···················· |
| E | 7506 | hCoV-19/India/GJ-GBRC260a/2020\|EPI_ISL_483851\|2020 | ···················· | ···················· | ····················· | ···················· | ···················· | ···A················ |
| E | 7507 | hCoV-19/India/GJ-GBRC260b/2020\|EPI_ISL_483852\|2020 | ···················· | ·A·················· | ····················· | ···················· | ···················· | ···················· |
| E | 7508 | hCoV-19/India/GJ-GBRC261a/2020\|EPI_ISL_483853\|2020 | ···················· | ···················· | ····················· | ···················· | ···················· | ···················· |
| E | 7509 | hCoV-19/India/GJ-GBRC261b/2020\|EPI_ISL_483854\|2020 | ···················· | ···················· | ····················· | ···················· | ···················· | ···A················ |
| E | 7510 | hCoV-19/India/GJ-GBRC262/2020\|EPI_ISL_483855\|2020- | ···················· | ···················· | ····················· | ···················· | ···················· | ···················· |
| E | 7511 | hCoV-19/India/GJ-GBRC263a/2020\|EPI_ISL_483856\|2020 | ···················· | ·A·················· | ····················· | ···················· | ···················· | ···A················ |
| E | 7512 | hCoV-19/India/GJ-GBRC263b/2020\|EPI_ISL_483857\|2020 | ···················· | ···················· | ····················· | ···················· | ···················· | ···················· |
| E | 7513 | hCoV-19/India/GJ-GBRC264a/2020\|EPI_ISL_483858\|2020 | ···················· | ···················· | ····················· | ···················· | ···················· | ···················· |
| E | 7514 | hCoV-19/India/GJ-GBRC264b/2020\|EPI_ISL_483859\|2020 | ···················· | ···················· | ····················· | ···················· | ···················· | ···A················ |
| E | 7515 | hCoV-19/India/GJ-GBRC265/2020\|EPI_ISL_483860\|2020- | ···················· | ···················· | ····················· | ···················· | ···················· | ···················· |
| E | 7516 | hCoV-19/India/GJ-GBRC266/2020\|EPI_ISL_483861\|2020- | ···················· | ···················· | ····················· | ···················· | ···················· | ···················· |
| E | 7517 | hCoV-19/India/GJ-GBRC267a/2020\|EPI_ISL_483862\|2020 | ···················· | ···················· | ····················· | ···················· | ···················· | ···················· |
| E | 7518 | hCoV-19/India/GJ-GBRC267b/2020\|EPI_ISL_483863\|2020 | ···················· | ·A·················· | ····················· | ···················· | ···················· | ···A················ |
| E | 7519 | hCoV-19/India/GJ-GBRC268a/2020\|EPI_ISL_483864\|2020 | ···················· | ···················· | ····················· | ···················· | ···················· | ···················· |
| E | 7520 | hCoV-19/India/GJ-GBRC268b/2020\|EPI_ISL_483865\|2020 | ···················· | ···················· | ····················· | ···················· | ···················· | ···A················ |
| E | 7521 | hCoV-19/India/GJ-GBRC269/2020\|EPI_ISL_483866\|2020- | ···················· | ···················· | ····················· | ···················· | ···················· | ···················· |
| E | 7522 | hCoV-19/India/GJ-GBRC270/2020\|EPI_ISL_483867\|2020- | ···················· | ···················· | ····················· | ···················· | ···················· | ···················· |
| E | 7523 | hCoV-19/India/GJ-GBRC271a/2020\|EPI_ISL_483868\|2020 | ···················· | ···················· | ····················· | ···················· | ···················· | ···A················ |
| E | 7524 | hCoV-19/India/GJ-GBRC271b/2020\|EPI_ISL_483869\|2020 | ···················· | ·A·················· | ····················· | ···················· | ···················· | ···················· |
| E | 7525 | hCoV-19/India/GJ-GBRC272a/2020\|EPI_ISL_483870\|2020 | ···················· | ···················· | ····················· | ···················· | ···················· | ···················· |
| E | 7526 | hCoV-19/India/GJ-GBRC272b/2020\|EPI_ISL_483871\|2020 | ···················· | ···················· | ····················· | ···················· | ···················· | ···A················ |
| E | 7527 | hCoV-19/India/GJ-GBRC273a/2020\|EPI_ISL_483872\|2020 | ···················· | ···················· | ····················· | ···················· | ···················· | ···················· |
| E | 7528 | hCoV-19/India/GJ-GBRC273b/2020\|EPI_ISL_483873\|2020 | ···················· | ···················· | ····················· | ···················· | ···················· | ···················· |
| E | 7529 | hCoV-19/England/BIRM-60718/2020\|EPI_ISL_483882\|202 | ···················· | ···················· | ····················· | ···················· | ···················· | ···················· |
| E | 7530 | hCoV-19/England/BIRM-60781/2020\|EPI_ISL_483887\|202 | ···················· | ···················· | ····················· | ···················· | ···················· | ···················· |
| E | 7531 | hCoV-19/England/BIRM-607DC/2020\|EPI_ISL_483889\|202 | ···················· | ···················· | ····················· | ···················· | ···················· | ···················· |
| E | 7532 | hCoV-19/England/BIRM-607FA/2020\|EPI_ISL_483890\|202 | ···················· | ···················· | ····················· | ···················· | ···················· | ···················· |
| E | 7533 | hCoV-19/England/BIRM-60806/2020\|EPI_ISL_483891\|202 | ···················· | ···················· | ····················· | ···················· | ···················· | ···················· |
| E | 7534 | hCoV-19/England/BIRM-60833/2020\|EPI_ISL_483893\|202 | ···················· | ···················· | ····················· | ···················· | ···················· | ···················· |
| E | 7535 | hCoV-19/England/BIRM-6088E/2020\|EPI_ISL_483896\|202 | ···················· | ···················· | ····················· | ···················· | ···················· | ···················· |
| E | 7536 | hCoV-19/England/BIRM-6089D/2020\|EPI_ISL_483897\|202 | ···················· | ···················· | ····················· | ···················· | ···················· | ···················· |
| E | 7537 | hCoV-19/England/BIRM-608BB/2020\|EPI_ISL_483899\|202 | ···················· | ···················· | ····················· | ···················· | ···················· | ···················· |
| E | 7538 | hCoV-19/Taiwan/NCKUH-005/2020\|EPI_ISL_493200\|2020- | ···················· | ···················· | ····················· | ···················· | ···················· | ···················· |
| E | 7539 | hCoV-19/Taiwan/NCKUH-007/2020\|EPI_ISL_493202\|2020- | ···················· | ···················· | ····················· | ···················· | ···················· | ···················· |
| E | 7540 | hCoV-19/Taiwan/NCKUH-012/2020\|EPI_ISL_493205\|2020- | ···················· | ···················· | ····················· | ···················· | ···················· | ···················· |
| E | 7541 | hCoV-19/Taiwan/NCKUH-013/2020\|EPI_ISL_493206\|2020- | ···················· | ···················· | ····················· | ···················· | ···················· | ···················· |
| E | 7542 | hCoV-19/Taiwan/NCKUH-014/2020\|EPI_ISL_493207\|2020- | ···················· | ···················· | ····················· | ···················· | ···················· | ···················· |
| E | 7543 | hCoV-19/USA/SC-USC-USC00173/2020\|EPI_ISL_493209\|20 | ···················· | ·········N·········· | ····················· | ···················· | ···················· | ···················· |
| E | 7544 | hCoV-19/USA/SC-USC-USC00437/2020\|EPI_ISL_493210\|20 | ···················· | ·········N·········· | ····················· | ···················· | ···················· | ···················· |
| E | 7545 | hCoV-19/USA/SC-USC-USC00433/2020\|EPI_ISL_493211\|20 | ···················· | ··············C····· | ····················· | ···················· | ···················· | ···················· |
| E | 7546 | hCoV-19/USA/SC-USC-PRG00427/2020\|EPI_ISL_493212\|20 | ···················· | ···················· | ····················· | ···················· | ···················· | ···················· |
| E | 7547 | hCoV-19/USA/CA-ALSR-1337/2020\|EPI_ISL_494500\|2020- | ···················· | ···················· | ····················· | ···················· | ···················· | ···················· |
| E | 7548 | hCoV-19/USA/CA-ALSR-1340/2020\|EPI_ISL_494502\|2020- | ···················· | ·A·················· | ····················· | ···················· | ···················· | ···················· |
| E | 7549 | hCoV-19/USA/CA-ALSR-1343/2020\|EPI_ISL_494503\|2020- | ···················· | ·A·················· | ····················· | ···················· | ···················· | ···················· |
| E | 7550 | hCoV-19/USA/AL-QDX-207/2020\|EPI_ISL_494504\|2020-05 | ···················· | ···················· | ····················· | ···················· | ···················· | ···················· |
| E | 7551 | hCoV-19/USA/CA-QDX-185/2020\|EPI_ISL_494505\|2020-04 | ···················· | ···················· | ····················· | ···················· | ···················· | ···················· |
| E | 7552 | hCoV-19/USA/CA-QDX-186/2020\|EPI_ISL_494506\|2020-04 | ···················· | ···················· | ····················· | ···················· | ···················· | ···················· |
| E | 7553 | hCoV-19/USA/CA-QDX-205/2020\|EPI_ISL_494507\|2020-05 | ···················· | ···················· | ····················· | ···················· | ···················· | ···················· |
| E | 7554 | hCoV-19/USA/CA-QDX-206/2020\|EPI_ISL_494508\|2020-05 | ···················· | ···················· | ····················· | ···················· | ···················· | ···················· |
| E | 7555 | hCoV-19/USA/CA-QDX-214/2020\|EPI_ISL_494509\|2020-05 | ···················· | ·A·················· | ····················· | ···················· | ···················· | ···················· |
| E | 7556 | hCoV-19/USA/GA-QDX-175/2020\|EPI_ISL_494510\|2020-04 | ···················· | ···················· | ····················· | ···················· | ···················· | ···················· |
| E | 7557 | hCoV-19/USA/GA-QDX-177/2020\|EPI_ISL_494511\|2020-04 | ···················· | ···················· | ····················· | ···················· | ···················· | ···················· |

| | | | | | | | |
|---|---|---|---|---|---|---|---|
| E | 7558 hCoV-19/USA/IL-QDX-176/2020\|EPI_ISL_494512\|2020-04 | ···················· | ···················· | ····················· | ···················· | ···················· | ···················· |
| E | 7559 hCoV-19/USA/IL-QDX-178/2020\|EPI_ISL_494513\|2020-04 | ···················· | ···················· | ····················· | ···················· | ···················· | ···················· |
| E | 7560 hCoV-19/USA/IL-QDX-195/2020\|EPI_ISL_494514\|2020-04 | ···················· | ···················· | ····················· | ···················· | ···················· | ···················· |
| E | 7561 hCoV-19/USA/IL-QDX-198/2020\|EPI_ISL_494515\|2020-04 | ···················· | ···················· | ····················· | ···················· | ···················· | ···················· |
| E | 7562 hCoV-19/USA/IL-QDX-199/2020\|EPI_ISL_494516\|2020-04 | ···················· | ···················· | ····················· | ···················· | ···················· | ···················· |
| E | 7563 hCoV-19/USA/IL-QDX-200/2020\|EPI_ISL_494517\|2020-04 | ···················· | ···················· | ····················· | ···················· | ···················· | ···················· |
| E | 7564 hCoV-19/USA/IN-QDX-182/2020\|EPI_ISL_494518\|2020-04 | ···················· | ···················· | ····················· | ···················· | ···················· | ···················· |
| E | 7565 hCoV-19/USA/IN-QDX-184/2020\|EPI_ISL_494519\|2020-04 | ···················· | ···················· | ····················· | ···················· | ···················· | ···················· |
| E | 7566 hCoV-19/USA/IN-QDX-192/2020\|EPI_ISL_494520\|2020-04 | ···················· | ···················· | ····················· | ···················· | ···················· | ···················· |
| E | 7567 hCoV-19/USA/LA-QDX-167/2020\|EPI_ISL_494521\|2020-04 | ···················· | ···················· | ····················· | ···················· | ···················· | ···················· |
| E | 7568 hCoV-19/USA/MI-QDX-183/2020\|EPI_ISL_494522\|2020-04 | ···················· | ···················· | ····················· | ···················· | ···················· | ···················· |
| E | 7569 hCoV-19/USA/NC-QDX-171/2020\|EPI_ISL_494523\|2020-04 | ···················· | ···················· | ····················· | ···················· | ···················· | ···················· |
| E | 7570 hCoV-19/USA/NC-QDX-172/2020\|EPI_ISL_494524\|2020-04 | ···················· | ···················· | ····················· | ···················· | ···················· | ···················· |
| E | 7571 hCoV-19/USA/NC-QDX-173/2020\|EPI_ISL_494525\|2020-04 | ···················· | ···················· | ····················· | ···················· | ···················· | ···················· |
| E | 7572 hCoV-19/USA/NC-QDX-174/2020\|EPI_ISL_494526\|2020-04 | ···················· | ···················· | ····················· | ···················· | ···················· | ···················· |
| E | 7573 hCoV-19/USA/NC-QDX-180/2020\|EPI_ISL_494527\|2020-04 | ···················· | ···················· | ····················· | ···················· | ···················· | ···················· |
| E | 7574 hCoV-19/USA/NC-QDX-181/2020\|EPI_ISL_494528\|2020-04 | ···················· | ···················· | ····················· | ···················· | ···················· | ···················· |
| E | 7575 hCoV-19/USA/NC-QDX-201/2020\|EPI_ISL_494529\|2020-04 | ···················· | ···················· | ····················· | ···················· | ···················· | ···················· |
| E | 7576 hCoV-19/USA/NC-QDX-202/2020\|EPI_ISL_494530\|2020-04 | ···················· | ···················· | ····················· | ···················· | ···················· | ···················· |
| E | 7577 hCoV-19/USA/NM-QDX-208/2020\|EPI_ISL_494531\|2020-04 | ···················· | ···················· | ····················· | ···················· | ···················· | ···················· |
| E | 7578 hCoV-19/USA/NM-QDX-209/2020\|EPI_ISL_494532\|2020-04 | ···················· | ···················· | ····················· | ···················· | ···················· | ···················· |
| E | 7579 hCoV-19/USA/NM-QDX-210/2020\|EPI_ISL_494533\|2020-04 | ···················· | ···················· | ····················· | ···················· | ···················· | ···················· |
| E | 7580 hCoV-19/USA/NM-QDX-211/2020\|EPI_ISL_494534\|2020-04 | ···················· | ···················· | ····················· | ···················· | ···················· | ···················· |
| E | 7581 hCoV-19/USA/NM-QDX-212/2020\|EPI_ISL_494535\|2020-04 | ···················· | ···················· | ····················· | ···················· | ···················· | ···················· |
| E | 7582 hCoV-19/USA/NM-QDX-213/2020\|EPI_ISL_494536\|2020-04 | ···················· | ···················· | ····················· | ···················· | ···················· | ···················· |
| E | 7583 hCoV-19/USA/NM-QDX-215/2020\|EPI_ISL_494537\|2020-05 | ···················· | ···················· | ····················· | ···················· | ···················· | ···················· |
| E | 7584 hCoV-19/USA/TX-QDX-166/2020\|EPI_ISL_494538\|2020-04 | ···················· | ···················· | ····················· | ···················· | ···················· | ···················· |
| E | 7585 hCoV-19/USA/TX-QDX-168/2020\|EPI_ISL_494539\|2020-04 | ···················· | ···················· | ····················· | ···················· | ···················· | ···················· |
| E | 7586 hCoV-19/USA/TX-QDX-169/2020\|EPI_ISL_494540\|2020-04 | ···················· | ···················· | ····················· | ···················· | ···················· | ···················· |
| E | 7587 hCoV-19/USA/TX-QDX-170/2020\|EPI_ISL_494541\|2020-04 | ···················· | ···················· | ····················· | ···················· | ···················· | ···················· |
| E | 7588 hCoV-19/USA/TX-QDX-187/2020\|EPI_ISL_494542\|2020-04 | ···················· | ···················· | ····················· | ···················· | ···················· | ···················· |
| E | 7589 hCoV-19/USA/TX-QDX-188/2020\|EPI_ISL_494543\|2020-04 | ···················· | ···················· | ····················· | ···················· | ···················· | ···················· |
| E | 7590 hCoV-19/USA/TX-QDX-189/2020\|EPI_ISL_494544\|2020-04 | ···················· | ···················· | ····················· | ···················· | ···················· | ···················· |
| E | 7591 hCoV-19/USA/TX-QDX-190/2020\|EPI_ISL_494545\|2020-04 | ···················· | ···················· | ····················· | ···················· | ···················· | ···················· |
| E | 7592 hCoV-19/USA/TX-QDX-191/2020\|EPI_ISL_494546\|2020-04 | ···················· | ···················· | ····················· | ···················· | ···················· | ···················· |
| E | 7593 hCoV-19/USA/TX-QDX-203/2020\|EPI_ISL_494547\|2020-04 | ···················· | ···················· | ····················· | ···················· | ···················· | ···················· |
| E | 7594 hCoV-19/USA/TX-QDX-204/2020\|EPI_ISL_494548\|2020-04 | ···················· | ···················· | ····················· | ···················· | ···················· | ···················· |
| E | 7595 hCoV-19/USA/WI-QDX-193/2020\|EPI_ISL_494549\|2020-04 | ···················· | ···················· | ····················· | ···················· | ···················· | ···················· |
| E | 7596 hCoV-19/USA/WI-QDX-194/2020\|EPI_ISL_494550\|2020-04 | ···················· | ···················· | ····················· | ···················· | ···················· | ···················· |
| E | 7597 hCoV-19/USA/WI-QDX-197/2020\|EPI_ISL_494552\|2020-04 | ···················· | ···················· | ····················· | ···················· | ···················· | ···················· |
| E | 7598 hCoV-19/USA/SC-USC-PRG467/2020\|EPI_ISL_494553\|2020 | ···················· | ···················· | ····················· | ···················· | ···················· | ···················· |
| E | 7599 hCoV-19/USA/SC-USC-PRG505/2020\|EPI_ISL_494554\|2020 | ···················· | ·········A·········· | ····················· | ···················· | ···················· | ···················· |
| E | 7600 hCoV-19/USA/SC-USC-PRG507/2020\|EPI_ISL_494555\|2020 | ···················· | ·········A·········· | ····················· | ···················· | ···················· | ···················· |
| E | 7601 hCoV-19/USA/CA-ALSR-1360/2020\|EPI_ISL_494568\|2020- | ···················· | ···················· | ····················· | ···················· | ···················· | ···················· |
| E | 7602 hCoV-19/USA/CA-ALSR-1362/2020\|EPI_ISL_494569\|2020- | ···················· | ···················· | ····················· | ···················· | ···················· | ···················· |
| E | 7603 hCoV-19/USA/CA-ALSR-1379/2020\|EPI_ISL_494570\|2020- | ···················· | ···················· | ····················· | ···················· | ···················· | ···················· |
| E | 7604 hCoV-19/USA/CA-ALSR-1380/2020\|EPI_ISL_494571\|2020- | ···················· | ···················· | ····················· | ···················· | ···················· | ···················· |
| E | 7605 hCoV-19/USA/CA-ALSR-1381/2020\|EPI_ISL_494572\|2020- | ···················· | ···················· | ····················· | ···················· | ···················· | ···················· |
| E | 7606 hCoV-19/USA/CA-ALSR-1391/2020\|EPI_ISL_494580\|2020- | ···················· | ·A·················· | ····················· | ···················· | ···················· | ···················· |
| E | 7607 hCoV-19/USA/CA-ALSR-1393/2020\|EPI_ISL_494581\|2020- | ···················· | ···················· | ····················· | ···················· | ···················· | ···················· |
| E | 7608 hCoV-19/USA/CA-ALSR-1397/2020\|EPI_ISL_494583\|2020- | ···················· | ·A·················· | ····················· | ···················· | ···················· | ···················· |
| E | 7609 hCoV-19/USA/CA-ALSR-1426/2020\|EPI_ISL_494595\|2020- | ···················· | ·A·················· | ····················· | ···················· | ···················· | ···················· |
| E | 7610 hCoV-19/USA/CA-ALSR-1428/2020\|EPI_ISL_494596\|2020- | ···················· | ···················· | ····················· | ···················· | ···················· | ···················· |
| E | 7611 hCoV-19/USA/CA-ALSR-1430/2020\|EPI_ISL_494597\|2020- | ···················· | ···················· | ····················· | ···················· | ···················· | ···················· |
| E | 7612 hCoV-19/USA/CA-ALSR-1432/2020\|EPI_ISL_494598\|2020- | ···················· | ···················· | ····················· | ···················· | ···················· | ···················· |
| E | 7613 hCoV-19/USA/WA-S1370/2020\|EPI_ISL_495820\|2020-05-1 | ···················· | ···················· | ····················· | ···················· | ···················· | ···················· |
| E | 7614 hCoV-19/USA/WA-S1371/2020\|EPI_ISL_495821\|2020-05-0 | ···················· | ···················· | ····················· | ···················· | ···················· | ···················· |
| E | 7615 hCoV-19/USA/WA-S1372/2020\|EPI_ISL_495822\|2020-05-1 | ···················· | ···················· | ····················· | ···················· | ···················· | ···················· |
| E | 7616 hCoV-19/USA/WA-S1373/2020\|EPI_ISL_495823\|2020-05-1 | ···················· | ···················· | ····················· | ···················· | ···················· | ···················· |
| E | 7617 hCoV-19/USA/WA-S1374/2020\|EPI_ISL_495824\|2020-05-1 | ···················· | ···················· | ····················· | ···················· | ···················· | ···················· |
| E | 7618 hCoV-19/USA/WA-S1375/2020\|EPI_ISL_495825\|2020-05-1 | ···················· | ···················· | ····················· | ···················· | ···················· | ···················· |
| E | 7619 hCoV-19/USA/WA-S1376/2020\|EPI_ISL_495826\|2020-05-1 | ···················· | ···················· | ····················· | ···················· | ···················· | ···················· |
| E | 7620 hCoV-19/USA/WA-S1377/2020\|EPI_ISL_495827\|2020-05-1 | ···················· | ···················· | ····················· | ···················· | ···················· | ···················· |
| E | 7621 hCoV-19/USA/WA-S1378/2020\|EPI_ISL_495828\|2020-05-1 | ···················· | ···················· | ····················· | ···················· | ···················· | ···················· |
| E | 7622 hCoV-19/USA/WA-S1379/2020\|EPI_ISL_495829\|2020-05-1 | ···················· | ···················· | ····················· | ···················· | ···················· | ···················· |
| E | 7623 hCoV-19/USA/WA-S1380/2020\|EPI_ISL_495830\|2020-05-1 | ···················· | ···················· | ····················· | ···················· | ···················· | ···················· |
| E | 7624 hCoV-19/USA/WA-S1381/2020\|EPI_ISL_495831\|2020-05-1 | ···················· | ···················· | ····················· | ···················· | ···················· | ···················· |
| E | 7625 hCoV-19/USA/WA-S1383/2020\|EPI_ISL_495833\|2020-05-1 | ···················· | ···················· | ····················· | ···················· | ···················· | ···················· |
| E | 7626 hCoV-19/USA/WA-S1384/2020\|EPI_ISL_495834\|2020-05-1 | ···················· | ···················· | ····················· | ···················· | ···················· | ···················· |
| E | 7627 hCoV-19/USA/WA-S1385/2020\|EPI_ISL_495835\|2020-05-1 | ···················· | ···················· | ····················· | ···················· | ···················· | ···················· |
| E | 7628 hCoV-19/USA/WA-S1386/2020\|EPI_ISL_495836\|2020-05-1 | ···················· | ···················· | ····················· | ···················· | ···················· | ···················· |
| E | 7629 hCoV-19/USA/WA-S1387/2020\|EPI_ISL_495837\|2020-05-1 | ···················· | ···················· | ····················· | ···················· | ···················· | ···················· |
| E | 7630 hCoV-19/USA/WA-S1388/2020\|EPI_ISL_495838\|2020-05-1 | ···················· | ···················· | ····················· | ···················· | ···················· | ···················· |
| E | 7631 hCoV-19/USA/WA-S1389/2020\|EPI_ISL_495839\|2020-05-1 | ···················· | ···················· | ····················· | ···················· | ···················· | ···················· |
| E | 7632 hCoV-19/USA/WA-S1390/2020\|EPI_ISL_495840\|2020-05-1 | ···················· | ···················· | ····················· | ···················· | ···················· | ···················· |
| E | 7633 hCoV-19/USA/WA-S1391/2020\|EPI_ISL_495841\|2020-05-1 | ···················· | ···················· | ····················· | ···················· | ···················· | ···················· |
| E | 7634 hCoV-19/USA/WA-S1392/2020\|EPI_ISL_495842\|2020-05-1 | ···················· | ···················· | ····················· | ···················· | ···················· | ···················· |
| E | 7635 hCoV-19/USA/WA-S1393/2020\|EPI_ISL_495843\|2020-05-1 | ···················· | ···················· | ····················· | ···················· | ···················· | ···················· |
| E | 7636 hCoV-19/USA/WA-S1394/2020\|EPI_ISL_495844\|2020-05-1 | ···················· | ···················· | ····················· | ···················· | ···················· | ···················· |
| E | 7637 hCoV-19/USA/WA-S1395/2020\|EPI_ISL_495845\|2020-05-1 | ···················· | ···················· | ····················· | ···················· | ···················· | ···················· |

```
E 7638 hCoV-19/USA/WA-S1397/2020|EPI_ISL_495847|2020-05-1 ···················· ···················· ····················· ···················· ···················· ····················
E 7639 hCoV-19/USA/WA-S1398/2020|EPI_ISL_495848|2020-05-1 ···················· ···················· ····················· ···················· ···················· ····················
E 7640 hCoV-19/USA/WA-S1399/2020|EPI_ISL_495849|2020-05-1 ···················· ···················· ····················· ···················· ···················· ····················
E 7641 hCoV-19/USA/WA-S1401/2020|EPI_ISL_495851|2020-05-1 ···················· ···················· ····················· ···················· ···················· ····················
E 7642 hCoV-19/USA/WA-S1402/2020|EPI_ISL_495852|2020-05-1 ···················· ···················· ····················· ···················· ···················· ····················
E 7643 hCoV-19/USA/WA-S1403/2020|EPI_ISL_495853|2020-05-1 ···················· ···················· ····················· ···················· ···················· ····················
E 7644 hCoV-19/USA/WA-S1404/2020|EPI_ISL_495854|2020-05-1 ···················· ···················· ····················· ···················· ···················· ····················
E 7645 hCoV-19/USA/WA-S1405/2020|EPI_ISL_495855|2020-05-1 ···················· ···················· ····················· ···················· ···················· ····················
E 7646 hCoV-19/USA/WA-S1406/2020|EPI_ISL_495856|2020-05-1 ···················· ···················· ····················· ···················· ···················· ····················
E 7647 hCoV-19/USA/WA-S1407/2020|EPI_ISL_495857|2020-05-1 ···················· ···················· ····················· ···················· ···················· ····················
E 7648 hCoV-19/USA/WA-S1408/2020|EPI_ISL_495858|2020-05-1 ···················· ···················· ····················· ···················· ···················· ····················
E 7649 hCoV-19/USA/WA-S1409/2020|EPI_ISL_495859|2020-05-1 ···················· ···················· ····················· ···················· ···················· ····················
E 7650 hCoV-19/USA/WA-S1410/2020|EPI_ISL_495860|2020-05-1 ···················· ···················· ····················· ···················· ···················· ····················
E 7651 hCoV-19/USA/WA-S1411/2020|EPI_ISL_495861|2020-05-1 ···················· ···················· ····················· ···················· ···················· ····················
E 7652 hCoV-19/USA/WA-S1412/2020|EPI_ISL_495862|2020-05-1 ···················· ···················· ····················· ···················· ···················· ····················
E 7653 hCoV-19/USA/WA-S1413/2020|EPI_ISL_495863|2020-05-2 ···················· ···················· ····················· ···················· ···················· ····················
E 7654 hCoV-19/USA/WA-S1414/2020|EPI_ISL_495864|2020-05-1 ···················· ···················· ····················· ···················· ···················· ····················
E 7655 hCoV-19/USA/WA-S1415/2020|EPI_ISL_495865|2020-05-1 ···················· ···················· ····················· ···················· ···················· ····················
E 7656 hCoV-19/USA/WA-S1416/2020|EPI_ISL_495866|2020-05-1 ···················· ···················· ····················· ···················· ···················· ····················
E 7657 hCoV-19/USA/WA-S1417/2020|EPI_ISL_495867|2020-05-1 ···················· ···················· ····················· ···················· ···················· ····················
E 7658 hCoV-19/USA/WA-S1418/2020|EPI_ISL_495868|2020-05-1 ···················· ···················· ····················· ···················· ···················· ····················
E 7659 hCoV-19/USA/WA-S1419/2020|EPI_ISL_495869|2020-05-1 ···················· ···················· ····················· ···················· ···················· ····················
E 7660 hCoV-19/USA/WA-S1424/2020|EPI_ISL_495874|2020-05-1 ···················· ···················· ····················· ···················· ···················· ····················
E 7661 hCoV-19/USA/WA-S1425/2020|EPI_ISL_495875|2020-05-2 ···················· ···················· ····················· ···················· ···················· ····················
E 7662 hCoV-19/USA/WA-S1426/2020|EPI_ISL_495876|2020-05-2 ···················· ···················· ····················· ···················· ···················· ····················
E 7663 hCoV-19/USA/WA-S1427/2020|EPI_ISL_495877|2020-05-2 ···················· ···················· ····················· ···················· ···················· ····················
E 7664 hCoV-19/USA/WA-S1428/2020|EPI_ISL_495878|2020-05-2 ···················· ···················· ····················· ···················· ···················· ····················
E 7665 hCoV-19/USA/WA-S1430/2020|EPI_ISL_495880|2020-05-2 ···················· ···················· ····················· ···················· ···················· ····················
E 7666 hCoV-19/USA/WA-S1431/2020|EPI_ISL_495881|2020-05-2 ···················· ···················· ····················· ···················· ···················· ····················
E 7667 hCoV-19/USA/WA-S1432/2020|EPI_ISL_495882|2020-05-2 ···················· ···················· ····················· ···················· ···················· ····················
E 7668 hCoV-19/USA/WA-S1433/2020|EPI_ISL_495883|2020-05-2 ···················· ···················· ····················· ···················· ···················· ····················
E 7669 hCoV-19/USA/WA-S1434/2020|EPI_ISL_495884|2020-05-2 ···················· ···················· ····················· ···················· ···················· ····················
E 7670 hCoV-19/USA/WA-S1439/2020|EPI_ISL_495889|2020-05-2 ···················· ···················· ····················· ···················· ···················· ····················
E 7671 hCoV-19/USA/WA-S1440/2020|EPI_ISL_495890|2020-05-2 ···················· ···················· ····················· ···················· ···················· ····················
E 7672 hCoV-19/USA/WA-S1441/2020|EPI_ISL_495891|2020-05-2 ···················· ···················· ····················· ···················· ···················· ····················
E 7673 hCoV-19/USA/WA-S1442/2020|EPI_ISL_495892|2020-05-2 ···················· ···················· ····················· ···················· ···················· ····················
E 7674 hCoV-19/USA/WA-S1443/2020|EPI_ISL_495893|2020-05-2 ···················· ···················· ····················· ···················· ···················· ····················
E 7675 hCoV-19/USA/WA-S1444/2020|EPI_ISL_495894|2020-05-2 ···················· ···················· ····················· ···················· ···················· ····················
E 7676 hCoV-19/USA/WA-S1445/2020|EPI_ISL_495895|2020-05-2 ···················· ···················· ····················· ···················· ···················· ····················
E 7677 hCoV-19/USA/WA-S1446/2020|EPI_ISL_495896|2020-05-2 ···················· ···················· ····················· ···················· ···················· ····················
E 7678 hCoV-19/USA/WA-S1447/2020|EPI_ISL_495897|2020-05-2 ···················· ···················· ····················· ···················· ···················· ····················
E 7679 hCoV-19/USA/WA-S1448/2020|EPI_ISL_495898|2020-05-2 ···················· ···················· ····················· ···················· ···················· ····················
E 7680 hCoV-19/USA/WA-S1449/2020|EPI_ISL_495899|2020-05-2 ···················· ···················· ····················· ···················· ···················· ····················
E 7681 hCoV-19/India/DL-NCDC1722-CSIR-IGIB/2020|EPI_ISL_4 ···················· ···················· ····················· ···················· ···················· ····················
E 7682 hCoV-19/India/DL-NCDC1733-CSIR-IGIB/2020|EPI_ISL_4 ···················· ···················· ····················· ···················· ···················· ····················
E 7683 hCoV-19/India/DL-NCDC1873-CSIR-IGIB/2020|EPI_ISL_4 ···················· ···················· ····················· ···················· ···················· ····················
E 7684 hCoV-19/India/DL-NCDC2188-CSIR-IGIB/2020|EPI_ISL_4 ···················· ·N·················· ····················· ···················· ···················· ····················
E 7685 hCoV-19/India/DL-NCDC2189-CSIR-IGIB/2020|EPI_ISL_4 ···················· ·R·················· ····················· ···················· ···················· ····················
E 7686 hCoV-19/India/DL-NCDC2192-CSIR-IGIB/2020|EPI_ISL_4 ···················· ·A·················· ····················· ···················· ···················· ···R················
E 7687 hCoV-19/Bangladesh/BCSIR-NILMRC-118/2020|EPI_ISL_4 ···················· ···················· ····················· ···················· ···················· ····················
E 7688 hCoV-19/Bangladesh/BCSIR-NILMRC-004-2/2020|EPI_ISL ···················· ···················· ····················· ···················· ···················· ····················
E 7689 hCoV-19/South Africa/KRISP-0175/2020|EPI_ISL_48270 ···················· ···················· ····················· ···················· ···················· ····················
E 7690 hCoV-19/South Africa/KRISP-0178/2020|EPI_ISL_48270 ···················· ···················· ····················· ···················· ···················· ····················
E 7691 hCoV-19/South Africa/KRISP-0179/2020|EPI_ISL_48270 ···················· ···················· ····················· ···················· ···················· ····················
E 7692 hCoV-19/South Africa/KRISP-0185/2020|EPI_ISL_48270 ···················· ···················· ····················· ···················· ···················· ····················
E 7693 hCoV-19/South Africa/KRISP-0190/2020|EPI_ISL_48270 ···················· ···················· ····················· ···················· ···················· ····················
E 7694 hCoV-19/India/GJ-GBRC239/2020|EPI_ISL_483820|2020- ···················· ···················· ····················· ···················· ···················· ····················
E 7695 hCoV-19/India/GJ-GBRC240/2020|EPI_ISL_483821|2020- ···················· ···················· ····················· ···················· ···················· ····················
E 7696 hCoV-19/India/GJ-GBRC241/2020|EPI_ISL_483822|2020- ···················· ···················· ····················· ···················· ···················· ····················
E 7697 hCoV-19/India/GJ-GBRC242/2020|EPI_ISL_483823|2020- ···················· ·A·················· ····················· ···················· ···················· ···A················
E 7698 hCoV-19/India/GJ-GBRC243/2020|EPI_ISL_483824|2020- ···················· ····A··············· ····················· ···················· ···················· ····················
E 7699 hCoV-19/India/GJ-GBRC244/2020|EPI_ISL_483825|2020- ···T················ ·A·················· ····················· ···················· ···················· ···A················
E 7700 hCoV-19/India/GJ-GBRC245/2020|EPI_ISL_483826|2020- ········T··········· ···················· ····················· ···················· ···················· ····················
E 7701 hCoV-19/India/GJ-GBRC246/2020|EPI_ISL_483827|2020- ···················· ···················· ····················· ···················· ···················· ····················
E 7702 hCoV-19/India/GJ-GBRC247/2020|EPI_ISL_483828|2020- ···················· ·A·················· ····················· ···················· ···················· ···A················
E 7703 hCoV-19/India/GJ-GBRC248a/2020|EPI_ISL_483829|2020 ········T··········· ····A··············· ····················· ···················· ···················· ····················
E 7704 hCoV-19/India/GJ-GBRC248b/2020|EPI_ISL_483830|2020 ········T··········· ····A··············· ····················· ···················· ···················· ····················
E 7705 hCoV-19/India/GJ-GBRC249a/2020|EPI_ISL_483831|2020 ···················· ·A·················· ····················· ···················· ···················· ···A················
E 7706 hCoV-19/India/GJ-GBRC249b/2020|EPI_ISL_483832|2020 ···················· ·A·················· ····················· ···················· ···················· ···A················
E 7707 hCoV-19/India/GJ-GBRC250a/2020|EPI_ISL_483833|2020 ···················· ·A·················· ····················· ···················· ···················· ···A················
E 7708 hCoV-19/India/GJ-GBRC250b/2020|EPI_ISL_483834|2020 ···················· ·A·················· ····················· ···················· ···················· ···A················
E 7709 hCoV-19/India/GJ-GBRC251a/2020|EPI_ISL_483835|2020 ···················· ···················· ····················· ···················· ···················· ····················
E 7710 hCoV-19/India/GJ-GBRC251b/2020|EPI_ISL_483836|2020 ···················· ···················· ····················· ···················· ···················· ····················
E 7711 hCoV-19/India/GJ-GBRC252/2020|EPI_ISL_483837|2020- ···················· ·A·················· ····················· ···················· ···················· ···A················
E 7712 hCoV-19/India/GJ-GBRC253a/2020|EPI_ISL_483838|2020 ···················· ·A·················· ····················· ···················· ···················· ···A················
E 7713 hCoV-19/India/GJ-GBRC253b/2020|EPI_ISL_483839|2020 ···················· ·A·················· ····················· ···················· ···················· ···A················
E 7714 hCoV-19/India/GJ-GBRC274/2020|EPI_ISL_483874|2020- ···················· ···················· ····················· ···················· ···················· ····················
E 7715 hCoV-19/India/GJ-GBRC275a/2020|EPI_ISL_483875|2020 ···················· ···················· ····················· ···················· ···················· ····················
E 7716 hCoV-19/India/GJ-GBRC275b/2020|EPI_ISL_483876|2020 ···················· ·A·················· ····················· ···················· ···················· ···A················
E 7717 hCoV-19/India/GJ-GBRC276/2020|EPI_ISL_483877|2020- ···················· ···················· ····················· ···················· ···A················ ····················
```

| | | | | | | | | |
|---|---|---|---|---|---|---|---|---|
| E | 7718 | hCoV-19/India/GJ-GBRC277a/2020\|EPI_ISL_483878\|2020 | ···················· | ···················· | ···················· | ···················· | ···················· | ···················· |
| E | 7719 | hCoV-19/India/GJ-GBRC277b/2020\|EPI_ISL_483879\|2020 | ···················· | ···················· | ···················· | ···················· | ···················· | ···A················ |
| E | 7720 | hCoV-19/USA/WA-UW-3024/2020\|EPI_ISL_486080\|2020-03 | ···················· | ···················· | ···················· | ···················· | ···················· | ···················· |
| E | 7721 | hCoV-19/USA/WA-UW-3026/2020\|EPI_ISL_486081\|2020-03 | ···················· | ···················· | ···················· | ···················· | ···················· | ···················· |
| E | 7722 | hCoV-19/USA/WA-UW-3030/2020\|EPI_ISL_486082\|2020-03 | ···················· | ···················· | ···················· | ···················· | ···················· | ···················· |
| E | 7723 | hCoV-19/USA/WA-UW-3041/2020\|EPI_ISL_486083\|2020-03 | ···················· | ···················· | ···················· | ···················· | ···················· | ···················· |
| E | 7724 | hCoV-19/USA/WA-UW-3046/2020\|EPI_ISL_486084\|2020-03 | ···················· | ···················· | ···················· | ···················· | ···················· | ···················· |
| E | 7725 | hCoV-19/USA/WA-UW-3048/2020\|EPI_ISL_486085\|2020-03 | ···················· | ···················· | ···················· | ···················· | ···················· | ···················· |
| E | 7726 | hCoV-19/USA/WA-UW-3052/2020\|EPI_ISL_486086\|2020-03 | ···················· | ···················· | ···················· | ···················· | ···················· | ···················· |
| E | 7727 | hCoV-19/USA/WA-UW-3060/2020\|EPI_ISL_486087\|2020-03 | ···················· | ···················· | ···················· | ···················· | ···················· | ···················· |
| E | 7728 | hCoV-19/USA/WA-UW-3069/2020\|EPI_ISL_486088\|2020-03 | ···················· | ···················· | ···················· | ···················· | ···················· | ···················· |
| E | 7729 | hCoV-19/USA/WA-UW-3070/2020\|EPI_ISL_486089\|2020-03 | ···················· | ···················· | ···················· | ···················· | ···················· | ···················· |
| E | 7730 | hCoV-19/USA/WA-UW-3078/2020\|EPI_ISL_486090\|2020-03 | ···················· | ···················· | ···················· | ···················· | ···················· | ···················· |
| E | 7731 | hCoV-19/USA/WA-UW-379/2020\|EPI_ISL_486093\|2020-03- | ···················· | ···················· | ···················· | ···················· | ···················· | ···················· |
| E | 7732 | hCoV-19/USA/WA-UW-3849/2020\|EPI_ISL_486094\|2020-03 | ···················· | ···················· | ···················· | ···················· | ···················· | ···················· |
| E | 7733 | hCoV-19/USA/WA-UW-4682/2020\|EPI_ISL_486097\|2020-04 | ···················· | ···················· | ···················· | ···················· | ···················· | ···················· |
| E | 7734 | hCoV-19/USA/WA-UW-4752/2020\|EPI_ISL_486098\|2020-03 | ···················· | ···················· | ···················· | ···················· | ···················· | ···················· |
| E | 7735 | hCoV-19/USA/WA-UW-4776/2020\|EPI_ISL_486099\|2020-03 | ···················· | ···················· | ···················· | ···················· | ···················· | ···················· |
| E | 7736 | hCoV-19/USA/CA-CZB-1909/2020\|EPI_ISL_486202\|2020-0 | ···················· | ···················· | ···················· | ···················· | ···················· | ···················· |
| E | 7737 | hCoV-19/USA/CA-CZB-1911/2020\|EPI_ISL_486204\|2020-0 | ···················· | ···················· | ···················· | ···················· | ···················· | ···················· |
| E | 7738 | hCoV-19/USA/CA-CZB-1912/2020\|EPI_ISL_486205\|2020-0 | ···················· | ···················· | ···················· | ···················· | ···················· | ···················· |
| E | 7739 | hCoV-19/USA/CA-CZB-1913/2020\|EPI_ISL_486206\|2020-0 | ···················· | ···················· | ···················· | ···················· | ···················· | ···················· |
| E | 7740 | hCoV-19/USA/CA-CZB-1915/2020\|EPI_ISL_486208\|2020-0 | ···················· | ···················· | ···················· | ···················· | ···················· | ···················· |
| E | 7741 | hCoV-19/USA/CA-CZB-1916/2020\|EPI_ISL_486209\|2020-0 | ···················· | ···················· | ···················· | ···················· | ···················· | ···················· |
| E | 7742 | hCoV-19/USA/CA-CZB-1918/2020\|EPI_ISL_486210\|2020-0 | ···················· | ···················· | ···················· | ···················· | ···················· | ···················· |
| E | 7743 | hCoV-19/USA/CA-CZB-1919/2020\|EPI_ISL_486211\|2020-0 | ···················· | ···················· | ···················· | ···················· | ···················· | ···················· |
| E | 7744 | hCoV-19/USA/CA-CZB-1920/2020\|EPI_ISL_486212\|2020-0 | ···················· | ···················· | ···················· | ···················· | ···················· | ···················· |
| E | 7745 | hCoV-19/USA/CA-CZB-1921/2020\|EPI_ISL_486213\|2020-0 | ···················· | ···················· | ···················· | ···················· | ···················· | ···················· |
| E | 7746 | hCoV-19/USA/CA-CZB-1922/2020\|EPI_ISL_486214\|2020-0 | ···················· | ···················· | ···················· | ···················· | ···················· | ···················· |
| E | 7747 | hCoV-19/USA/CA-CZB-1923/2020\|EPI_ISL_486215\|2020-0 | ···················· | ···················· | ···················· | ···················· | ···················· | ···················· |
| E | 7748 | hCoV-19/USA/CA-CZB-1926/2020\|EPI_ISL_486216\|2020-0 | ···················· | ···················· | ···················· | ···················· | ···················· | ···················· |
| E | 7749 | hCoV-19/USA/CA-CZB-1928/2020\|EPI_ISL_486217\|2020-0 | ···················· | ···················· | ···················· | ···················· | ···················· | ···················· |
| E | 7750 | hCoV-19/USA/CA-CZB-1929/2020\|EPI_ISL_486218\|2020-0 | ···················· | ···················· | ···················· | ···················· | ···················· | ···················· |
| E | 7751 | hCoV-19/USA/CA-CZB-1931/2020\|EPI_ISL_486220\|2020-0 | ···················· | ···················· | ···················· | ···················· | ···················· | ···················· |
| E | 7752 | hCoV-19/USA/CA-CZB-1932/2020\|EPI_ISL_486221\|2020-0 | ···················· | ···················· | ···················· | ···················· | ···················· | ···················· |
| E | 7753 | hCoV-19/USA/CA-CZB-1944/2020\|EPI_ISL_486222\|2020-0 | ···T················ | ···················· | ···················· | ···················· | ···················· | ···················· |
| E | 7754 | hCoV-19/USA/CA-CZB-1959/2020\|EPI_ISL_486231\|2020-0 | ···················· | ···················· | ···················· | ···················· | ···················· | ···················· |
| E | 7755 | hCoV-19/USA/CA-CZB-1960/2020\|EPI_ISL_486232\|2020-0 | ···················· | ···················· | ···················· | ···················· | ···················· | ···················· |
| E | 7756 | hCoV-19/USA/CA-CZB-1961/2020\|EPI_ISL_486233\|2020-0 | ···················· | ···················· | ···················· | ···················· | ···················· | ···················· |
| E | 7757 | hCoV-19/USA/CA-CZB-1962/2020\|EPI_ISL_486234\|2020-0 | ···················· | ···················· | ···················· | ···················· | ···················· | ···················· |
| E | 7758 | hCoV-19/USA/CA-CZB-1963/2020\|EPI_ISL_486235\|2020-0 | ···················· | ···················· | ···················· | ···················· | ···················· | ···················· |
| E | 7759 | hCoV-19/USA/CA-CZB-1964/2020\|EPI_ISL_486236\|2020-0 | ···················· | ···················· | ···················· | ···················· | ···················· | ···················· |
| E | 7760 | hCoV-19/USA/CA-CZB-1965/2020\|EPI_ISL_486237\|2020-0 | ···················· | ···················· | ···················· | ···················· | ···················· | ···················· |
| E | 7761 | hCoV-19/USA/CA-CZB-1966/2020\|EPI_ISL_486238\|2020-0 | ···················· | ···················· | ···················· | ···················· | ···················· | ···················· |
| E | 7762 | hCoV-19/USA/CA-CZB-1967/2020\|EPI_ISL_486239\|2020-0 | ···················· | ···················· | ···················· | ···················· | ···················· | ···················· |
| E | 7763 | hCoV-19/USA/CA-CZB-1968/2020\|EPI_ISL_486240\|2020-0 | ···················· | ···················· | ···················· | ···················· | ···················· | ···················· |
| E | 7764 | hCoV-19/USA/CA-CZB-1969/2020\|EPI_ISL_486241\|2020-0 | ···················· | ···················· | ···················· | ···················· | ···················· | ···················· |
| E | 7765 | hCoV-19/USA/CA-CZB-1970/2020\|EPI_ISL_486242\|2020-0 | ···················· | ···················· | ···················· | ···················· | ···················· | ···················· |
| E | 7766 | hCoV-19/USA/CA-CZB-1971/2020\|EPI_ISL_486243\|2020-0 | ···················· | ···················· | ···················· | ···················· | ···················· | ···················· |
| E | 7767 | hCoV-19/USA/CA-CZB-1972/2020\|EPI_ISL_486244\|2020-0 | ···T················ | ···················· | ···················· | ···················· | ···················· | ···················· |
| E | 7768 | hCoV-19/USA/CA-CZB-1978/2020\|EPI_ISL_486247\|2020-0 | ···················· | ···················· | ···················· | ···················· | ···················· | ···················· |
| E | 7769 | hCoV-19/USA/CA-CZB-1980/2020\|EPI_ISL_486249\|2020-0 | ···················· | ···················· | ···················· | ···················· | ···················· | ···················· |
| E | 7770 | hCoV-19/USA/CA-CZB-1982/2020\|EPI_ISL_486250\|2020-0 | ···················· | ···················· | ···················· | ···················· | ···················· | ···················· |
| E | 7771 | hCoV-19/USA/CA-CZB-1984/2020\|EPI_ISL_486251\|2020-0 | ···················· | ···················· | ···················· | ···················· | ···················· | ···················· |
| E | 7772 | hCoV-19/USA/CA-CZB-1985/2020\|EPI_ISL_486252\|2020-0 | ···················· | ···················· | ···················· | ···················· | ···················· | ···················· |
| E | 7773 | hCoV-19/USA/CA-CZB-1986/2020\|EPI_ISL_486253\|2020-0 | ···················· | ···················· | ···················· | ···················· | ···················· | ···················· |
| E | 7774 | hCoV-19/USA/CA-CZB-1992/2020\|EPI_ISL_486255\|2020-0 | ···················· | ···················· | ···················· | ···················· | ···················· | ···················· |
| E | 7775 | hCoV-19/USA/CA-CZB-1993/2020\|EPI_ISL_486256\|2020-0 | ···················· | ···················· | ···················· | ···················· | ···················· | ···················· |
| E | 7776 | hCoV-19/USA/CA-CZB-1994/2020\|EPI_ISL_486257\|2020-0 | ···················· | ···················· | ···················· | ···················· | ···················· | ···················· |
| E | 7777 | hCoV-19/USA/CA-CZB-1995/2020\|EPI_ISL_486258\|2020-0 | ···················· | ···················· | ···················· | ···················· | ···················· | ···················· |
| E | 7778 | hCoV-19/USA/CA-CZB-1999/2020\|EPI_ISL_486259\|2020-0 | ···················· | ···················· | ···················· | ···················· | ···················· | ···················· |
| E | 7779 | hCoV-19/USA/CA-CZB-2000/2020\|EPI_ISL_486260\|2020-0 | ···················· | ···················· | ···················· | ···················· | ···················· | ···················· |
| E | 7780 | hCoV-19/USA/CA-CZB-2001/2020\|EPI_ISL_486261\|2020-0 | ···················· | ···················· | ···················· | ···················· | ···················· | ···················· |
| E | 7781 | hCoV-19/USA/CA-CZB-2002/2020\|EPI_ISL_486262\|2020-0 | ···················· | ···················· | ···················· | ···················· | ···················· | ···················· |
| E | 7782 | hCoV-19/USA/CA-CZB-2003/2020\|EPI_ISL_486263\|2020-0 | ···················· | ···················· | ···················· | ···················· | ···················· | ···················· |
| E | 7783 | hCoV-19/USA/CA-CZB-2004/2020\|EPI_ISL_486264\|2020-0 | ···················· | ···················· | ···················· | ···················· | ···················· | ···················· |
| E | 7784 | hCoV-19/USA/CA-CZB-2006/2020\|EPI_ISL_486265\|2020-0 | ···················· | ···················· | ···················· | ···················· | ···················· | ···················· |
| E | 7785 | hCoV-19/USA/CA-CZB-2011/2020\|EPI_ISL_486266\|2020-0 | ···················· | ···················· | ···················· | ···················· | ···················· | ···················· |
| E | 7786 | hCoV-19/USA/CA-CZB-2013/2020\|EPI_ISL_486267\|2020-0 | ···················· | ···················· | ···················· | ···················· | ···················· | ···················· |
| E | 7787 | hCoV-19/USA/CA-CZB-2015/2020\|EPI_ISL_486268\|2020-0 | ···················· | ···················· | ···················· | ···················· | ···················· | ···················· |
| E | 7788 | hCoV-19/USA/CA-CZB-2016/2020\|EPI_ISL_486269\|2020-0 | ···T················ | ···················· | ···················· | ···················· | ···················· | ···················· |
| E | 7789 | hCoV-19/USA/CA-CZB-2017/2020\|EPI_ISL_486270\|2020-0 | ···················· | ···················· | ···················· | ···················· | ···················· | ···················· |
| E | 7790 | hCoV-19/USA/CA-CZB-2018/2020\|EPI_ISL_486271\|2020-0 | ···················· | ···················· | ···················· | ···················· | ···················· | ···················· |
| E | 7791 | hCoV-19/USA/CA-CZB-2020/2020\|EPI_ISL_486272\|2020-0 | ···················· | ···················· | ···················· | ···················· | ···················· | ···················· |
| E | 7792 | hCoV-19/USA/CA-CZB-2022/2020\|EPI_ISL_486274\|2020-0 | ···················· | ···················· | ···················· | ···················· | ···················· | ···················· |
| E | 7793 | hCoV-19/USA/CA-CZB-2023/2020\|EPI_ISL_486275\|2020-0 | ···················· | ···················· | ···················· | ···················· | ···················· | ···················· |
| E | 7794 | hCoV-19/USA/CA-CZB-2024/2020\|EPI_ISL_486276\|2020-0 | ···················· | ···················· | ···················· | ···················· | ···················· | ···················· |
| E | 7795 | hCoV-19/USA/CA-CZB-2030/2020\|EPI_ISL_486278\|2020-0 | ···················· | ···················· | ···················· | ···················· | ···················· | ···················· |
| E | 7796 | hCoV-19/USA/CA-CZB-2031/2020\|EPI_ISL_486279\|2020-0 | ···················· | ···················· | ···················· | ···················· | ···················· | ···················· |
| E | 7797 | hCoV-19/USA/CA-CZB-2035/2020\|EPI_ISL_486281\|2020-0 | ···················· | ···················· | ···················· | ···················· | ···················· | ···················· |

| | | | | | | | | |
|---|---|---|---|---|---|---|---|---|
| E | 7798 | hCoV-19/USA/CA-CZB-2036/2020\|EPI_ISL_486282\|2020-0 | ···················· | ···················· | ···················· | ···················· | ···················· | ···················· |
| E | 7799 | hCoV-19/USA/CA-CZB-2037/2020\|EPI_ISL_486283\|2020-0 | ···················· | ···················· | ···················· | ···················· | ···················· | ···················· |
| E | 7800 | hCoV-19/USA/CA-CZB-2039/2020\|EPI_ISL_486284\|2020-0 | ···················· | ···················· | ···················· | ···················· | ···················· | ···················· |
| E | 7801 | hCoV-19/USA/CA-CZB-2041/2020\|EPI_ISL_486285\|2020-0 | ···················· | ···················· | ···················· | ···················· | ···················· | ···················· |
| E | 7802 | hCoV-19/USA/CA-CZB-2042/2020\|EPI_ISL_486286\|2020-0 | ···················· | ···················· | ···················· | ···················· | ···················· | ···················· |
| E | 7803 | hCoV-19/USA/CA-CZB-2043/2020\|EPI_ISL_486287\|2020-0 | ···················· | ···················· | ···················· | ···················· | ···················· | ···················· |
| E | 7804 | hCoV-19/USA/CA-CZB-2044/2020\|EPI_ISL_486288\|2020-0 | ···················· | ···················· | ···················· | ···················· | ···················· | ···················· |
| E | 7805 | hCoV-19/USA/CA-CZB-2045/2020\|EPI_ISL_486289\|2020-0 | ···················· | ···················· | ···················· | ···················· | ···················· | ···················· |
| E | 7806 | hCoV-19/USA/CA-CZB-2046/2020\|EPI_ISL_486290\|2020-0 | ···················· | ···················· | ···················· | ···················· | ···················· | ···················· |
| E | 7807 | hCoV-19/USA/CA-CZB-2047/2020\|EPI_ISL_486291\|2020-0 | ···················· | ···················· | ···················· | ···················· | ···················· | ···················· |
| E | 7808 | hCoV-19/USA/CA-CZB-2049/2020\|EPI_ISL_486293\|2020-0 | ···················· | ···················· | ···················· | ···················· | ···················· | ···················· |
| E | 7809 | hCoV-19/USA/CA-CZB-2050/2020\|EPI_ISL_486294\|2020-0 | ···················· | ···················· | ···················· | ···················· | ···················· | ···················· |
| E | 7810 | hCoV-19/USA/CA-CZB-2051/2020\|EPI_ISL_486295\|2020-0 | ···················· | ···················· | ···················· | ···················· | ···················· | ···················· |
| E | 7811 | hCoV-19/USA/CA-CZB-2052/2020\|EPI_ISL_486296\|2020-0 | ···················· | ···················· | ···················· | ···················· | ···················· | ···················· |
| E | 7812 | hCoV-19/USA/CA-CZB-2053/2020\|EPI_ISL_486297\|2020-0 | ···················· | ···················· | ···················· | ···················· | ···················· | ···················· |
| E | 7813 | hCoV-19/USA/CA-CZB-2054/2020\|EPI_ISL_486298\|2020-0 | ···················· | ···················· | ···················· | ···················· | ···················· | ···················· |
| E | 7814 | hCoV-19/USA/CA-CZB-2056/2020\|EPI_ISL_486299\|2020-0 | ···················· | ···················· | ···················· | ···················· | ···················· | ···················· |
| E | 7815 | hCoV-19/England/BRIS-12B5C3/2020\|EPI_ISL_487524\|20 | ···················· | ···················· | ···················· | ···················· | ···················· | ···················· |
| E | 7816 | hCoV-19/England/LOND-D6513/2020\|EPI_ISL_487525\|202 | ···················· | ···················· | ···················· | ···················· | ···················· | ···················· |
| E | 7817 | hCoV-19/England/BRIS-12B815/2020\|EPI_ISL_487526\|20 | ···················· | ···················· | ···················· | ···················· | ···················· | ···················· |
| E | 7818 | hCoV-19/England/BRIS-12B2BD/2020\|EPI_ISL_487527\|20 | ···················· | ···················· | ···················· | ···················· | ···················· | ···················· |
| E | 7819 | hCoV-19/England/BRIS-12B6A2/2020\|EPI_ISL_487528\|20 | ···················· | ···················· | ···················· | ···················· | ···················· | ···················· |
| E | 7820 | hCoV-19/England/LOND-D64F8/2020\|EPI_ISL_487529\|202 | ···················· | ···················· | ···················· | ···················· | ···················· | ···················· |
| E | 7821 | hCoV-19/England/BRIS-12B648/2020\|EPI_ISL_487530\|20 | ···················· | ···················· | ···················· | ···················· | ···················· | ···················· |
| E | 7822 | hCoV-19/England/BRIS-12B39C/2020\|EPI_ISL_487531\|20 | ···················· | ···················· | ···················· | ···················· | ···················· | ···················· |
| E | 7823 | hCoV-19/England/BRIS-12B2CC/2020\|EPI_ISL_487532\|20 | ···················· | ···················· | ···················· | ···················· | ···················· | ···················· |
| E | 7824 | hCoV-19/England/BRIS-12B2AE/2020\|EPI_ISL_487533\|20 | ···················· | ···················· | ···················· | ···················· | ···················· | ···················· |
| E | 7825 | hCoV-19/England/BRIS-12B6DF/2020\|EPI_ISL_487535\|20 | ···················· | ···················· | ···················· | ···················· | ···················· | ···················· |
| E | 7826 | hCoV-19/England/BRIS-12B772/2020\|EPI_ISL_487536\|20 | ···················· | ···················· | ···················· | ···················· | ···················· | ···················· |
| E | 7827 | hCoV-19/England/LOND-D6391/2020\|EPI_ISL_487537\|202 | ···················· | ···················· | ···················· | ···················· | ···················· | ···················· |
| E | 7828 | hCoV-19/England/BRIS-12B411/2020\|EPI_ISL_487538\|20 | ···················· | ···················· | ···················· | ···················· | ···················· | ···················· |
| E | 7829 | hCoV-19/England/BRIS-12B718/2020\|EPI_ISL_487539\|20 | ···················· | ···················· | ···················· | ···················· | ···················· | ···················· |
| E | 7830 | hCoV-19/England/LOND-D6452/2020\|EPI_ISL_487540\|202 | ···················· | ···················· | ···················· | ···················· | ···················· | ···················· |
| E | 7831 | hCoV-19/England/BRIS-12B842/2020\|EPI_ISL_487541\|20 | ···················· | ···················· | ···················· | ···················· | ···················· | ···················· |
| E | 7832 | hCoV-19/England/BRIS-12B5A5/2020\|EPI_ISL_487542\|20 | ···················· | ···················· | ···················· | ···················· | ···················· | ···················· |
| E | 7833 | hCoV-19/England/BRIS-12B48A/2020\|EPI_ISL_487543\|20 | ···················· | ···················· | ···················· | ···················· | ···················· | ···················· |
| E | 7834 | hCoV-19/England/BRIS-12B44E/2020\|EPI_ISL_487544\|20 | ···················· | ···················· | ···················· | ···················· | ···················· | ···················· |
| E | 7835 | hCoV-19/England/BRIS-12B3BA/2020\|EPI_ISL_487545\|20 | ···················· | ···················· | ···················· | ···················· | ···················· | ···················· |
| E | 7836 | hCoV-19/England/BRIS-12B36F/2020\|EPI_ISL_487547\|20 | ···················· | ···················· | ···················· | ···················· | ···················· | ···················· |
| E | 7837 | hCoV-19/England/BRIS-12B5E1/2020\|EPI_ISL_487549\|20 | ···················· | ···················· | ···················· | ···················· | ···················· | ···················· |
| E | 7838 | hCoV-19/England/LOND-D6531/2020\|EPI_ISL_487550\|202 | ···················· | ···················· | ···················· | ···················· | ···················· | ···················· |
| E | 7839 | hCoV-19/England/BRIS-12B4E4/2020\|EPI_ISL_487551\|20 | ···················· | ···A················ | ···················· | ···················· | ···················· | ···················· |
| E | 7840 | hCoV-19/England/BRIS-12B46C/2020\|EPI_ISL_487552\|20 | ···················· | ···················· | ···················· | ···················· | ···················· | ···················· |
| E | 7841 | hCoV-19/England/BRIS-12B806/2020\|EPI_ISL_487553\|20 | ···················· | ···················· | ···················· | ···················· | ···················· | ···················· |
| E | 7842 | hCoV-19/England/BRIS-12B781/2020\|EPI_ISL_487554\|20 | ···················· | ···················· | ···················· | ···················· | ···················· | ···················· |
| E | 7843 | hCoV-19/England/BRIS-12B4F3/2020\|EPI_ISL_487561\|20 | ···················· | ···A················ | ···················· | ···················· | ···················· | ···················· |
| E | 7844 | hCoV-19/England/LOND-D63EC/2020\|EPI_ISL_487562\|202 | ···················· | ···················· | ···················· | ···················· | ···················· | ···················· |
| E | 7845 | hCoV-19/England/BRIS-12B790/2020\|EPI_ISL_487563\|20 | ···················· | ···················· | ···················· | ···················· | ···················· | ···················· |
| E | 7846 | hCoV-19/England/LOND-D6470/2020\|EPI_ISL_487564\|202 | ···················· | ···················· | ···················· | ···················· | ···················· | ···················· |
| E | 7847 | hCoV-19/England/BRIS-12B5B4/2020\|EPI_ISL_487565\|20 | ···················· | ···················· | ···················· | ···················· | ···················· | ···················· |
| E | 7848 | hCoV-19/England/BRIS-12B727/2020\|EPI_ISL_487566\|20 | ···················· | ·A·················· | ···················· | ···················· | ···················· | ···················· |
| E | 7849 | hCoV-19/England/BRIS-12B666/2020\|EPI_ISL_487567\|20 | ···················· | ···················· | ···················· | ···················· | ···················· | ···················· |
| E | 7850 | hCoV-19/England/LOND-D6443/2020\|EPI_ISL_487568\|202 | ···················· | ···················· | ···················· | ···················· | ···················· | ···················· |
| E | 7851 | hCoV-19/England/BRIS-12B3C9/2020\|EPI_ISL_487570\|20 | ···················· | ···················· | ···················· | ···················· | ···················· | ···················· |
| E | 7852 | hCoV-19/England/BRIS-12B47B/2020\|EPI_ISL_487571\|20 | ···················· | ···················· | ···················· | ···················· | ···················· | ···················· |
| E | 7853 | hCoV-19/England/LOND-D662F/2020\|EPI_ISL_487572\|202 | ···················· | ···················· | ···················· | ···················· | ···················· | ···················· |
| E | 7854 | hCoV-19/England/BRIS-12B736/2020\|EPI_ISL_487573\|20 | ···················· | ···················· | ···················· | ···················· | ···················· | ···················· |
| E | 7855 | hCoV-19/England/BRIS-12B596/2020\|EPI_ISL_487574\|20 | ···················· | ···················· | ···················· | ···················· | ···················· | ···················· |
| E | 7856 | hCoV-19/England/BRIS-12B305/2020\|EPI_ISL_487575\|20 | ···················· | ···················· | ···················· | ···················· | ···················· | ···················· |
| E | 7857 | hCoV-19/England/BRIS-12B7AF/2020\|EPI_ISL_487576\|20 | ···················· | ···················· | ···················· | ···················· | ···················· | ···················· |
| E | 7858 | hCoV-19/England/BRIS-12B350/2020\|EPI_ISL_487577\|20 | ···················· | ···················· | ··T················· | ···················· | ···················· | ···················· |
| E | 7859 | hCoV-19/England/LOND-D656E/2020\|EPI_ISL_487578\|202 | ···················· | ···················· | ···················· | ···················· | ···················· | ···················· |
| E | 7860 | hCoV-19/England/BRIS-12B62A/2020\|EPI_ISL_487579\|20 | ···················· | ···················· | ···················· | ···················· | ···················· | ···················· |
| E | 7861 | hCoV-19/England/BRIS-12B684/2020\|EPI_ISL_487580\|20 | ···················· | ···················· | ···················· | ···················· | ···················· | ···················· |
| E | 7862 | hCoV-19/England/LOND-D64E9/2020\|EPI_ISL_487581\|202 | ···················· | ···················· | ···················· | ···················· | ···················· | ···················· |
| E | 7863 | hCoV-19/England/LOND-D655F/2020\|EPI_ISL_487582\|202 | ···················· | ···················· | ···················· | ···················· | ···················· | ···················· |
| E | 7864 | hCoV-19/England/BRIS-12B61B/2020\|EPI_ISL_487583\|20 | ···················· | ···················· | ···················· | ···················· | ···················· | ···················· |
| E | 7865 | hCoV-19/England/LOND-D663E/2020\|EPI_ISL_487584\|202 | ···················· | ···················· | ···················· | ···················· | ···················· | ···················· |
| E | 7866 | hCoV-19/England/LOND-D648F/2020\|EPI_ISL_487585\|202 | ···················· | ···················· | ···················· | ···················· | ···················· | ···················· |
| E | 7867 | hCoV-19/England/BRIS-12B763/2020\|EPI_ISL_487586\|20 | ···················· | ···················· | ···················· | ···················· | ···················· | ···················· |
| E | 7868 | hCoV-19/England/BRIS-12B53C/2020\|EPI_ISL_487587\|20 | ···················· | ···················· | ···················· | ···················· | ···················· | ···················· |
| E | 7869 | hCoV-19/England/BRIS-12B54B/2020\|EPI_ISL_487588\|20 | ···················· | ···················· | ···················· | ···················· | ···················· | ···················· |
| E | 7870 | hCoV-19/England/LOND-D6407/2020\|EPI_ISL_487589\|202 | ···················· | ···················· | ···················· | ···················· | ···················· | ···················· |
| E | 7871 | hCoV-19/England/BRIS-12B5F0/2020\|EPI_ISL_487590\|20 | ···················· | ···················· | ···················· | ···················· | ···················· | ···················· |
| E | 7872 | hCoV-19/England/BRIS-12B7EB/2020\|EPI_ISL_487591\|20 | ···················· | ···················· | ···················· | ···················· | ···················· | ···················· |
| E | 7873 | hCoV-19/England/BRIS-12B3F6/2020\|EPI_ISL_487592\|20 | ···················· | ···················· | ···················· | ···················· | ···················· | ···················· |
| E | 7874 | hCoV-19/England/BRIS-12B675/2020\|EPI_ISL_487594\|20 | ···················· | ···················· | ···················· | ···················· | ···················· | ···················· |
| E | 7875 | hCoV-19/England/LOND-D659B/2020\|EPI_ISL_487595\|202 | ···················· | ···················· | ···················· | ···················· | ···················· | ···················· |
| E | 7876 | hCoV-19/England/BRIS-12B52D/2020\|EPI_ISL_487596\|20 | ···················· | ···················· | ···················· | ···················· | ···················· | ···················· |
| E | 7877 | hCoV-19/England/LOND-D64CB/2020\|EPI_ISL_487597\|202 | ···················· | ···················· | ···················· | ···················· | ···················· | ···················· |

| | | | | | | | | |
|---|---|---|---|---|---|---|---|---|
| E | 7878 hCoV-19/England/BRIS-12B639/2020\|EPI_ISL_487598\|20 | ···················· | ···················· | ···················· | ···················· | ···················· | ···················· | |
| E | 7879 hCoV-19/England/BRIS-12B50F/2020\|EPI_ISL_487599\|20 | ···················· | ·A·················· | ···················· | ···················· | ···················· | ···················· | |
| E | 7880 hCoV-19/England/WSFT-25C17A1/2020\|EPI_ISL_488854\|2 | ···················· | ···················· | ···················· | ···················· | ···················· | ···················· | |
| E | 7881 hCoV-19/England/WSFT-25C1A5D/2020\|EPI_ISL_488856\|2 | ···················· | ···················· | ···················· | ···················· | ···················· | ···················· | |
| E | 7882 hCoV-19/England/HECH-259A2A0/2020\|EPI_ISL_488857\|2 | ···················· | ···················· | ···················· | ···················· | ···················· | ···················· | |
| E | 7883 hCoV-19/England/HECH-259A219/2020\|EPI_ISL_488858\|2 | ···················· | ···················· | ···················· | ···················· | ···················· | ···················· | |
| E | 7884 hCoV-19/England/HECH-259A2FB/2020\|EPI_ISL_488859\|2 | ···················· | ···················· | ···················· | ·······C············ | ···················· | ···················· | |
| E | 7885 hCoV-19/England/HECH-259A1FE/2020\|EPI_ISL_488860\|2 | ···················· | ···················· | ···················· | ·······C············ | ···················· | ···················· | |
| E | 7886 hCoV-19/England/WSFT-25C1A11/2020\|EPI_ISL_488861\|2 | ···················· | ···················· | ···················· | ···················· | ···················· | ···················· | |
| E | 7887 hCoV-19/England/HECH-259A1B2/2020\|EPI_ISL_488862\|2 | ···················· | ···················· | ···················· | ···················· | ···················· | ···················· | |
| E | 7888 hCoV-19/England/WSFT-25C18EA/2020\|EPI_ISL_488863\|2 | ···················· | ···················· | ···················· | ···················· | ···················· | ···················· | |
| E | 7889 hCoV-19/England/HECH-259A264/2020\|EPI_ISL_488865\|2 | ···················· | ···················· | ···················· | ···················· | ···················· | ···················· | |
| E | 7890 hCoV-19/England/HECH-259A2BF/2020\|EPI_ISL_488866\|2 | ···················· | ···················· | ···················· | ·······C············ | ···················· | ···················· | |
| E | 7891 hCoV-19/England/HECH-259A255/2020\|EPI_ISL_488867\|2 | ···················· | ···················· | ···················· | ···················· | ···················· | ···················· | |
| E | 7892 hCoV-19/England/HECH-259A1EF/2020\|EPI_ISL_488868\|2 | ···················· | ···················· | ···················· | ·······C············ | ···················· | ···················· | |
| E | 7893 hCoV-19/England/HECH-259A282/2020\|EPI_ISL_488869\|2 | ···················· | ···················· | ···················· | ···················· | ···················· | ···················· | |
| E | 7894 hCoV-19/England/HECH-259A316/2020\|EPI_ISL_488870\|2 | ···················· | ···················· | ···················· | ···················· | ···················· | ···················· | |
| E | 7895 hCoV-19/England/HECH-259A0F1/2020\|EPI_ISL_488871\|2 | ···················· | ···················· | ···················· | ···················· | ···················· | ···················· | |
| E | 7896 hCoV-19/England/WSFT-25C17ED/2020\|EPI_ISL_488872\|2 | ···················· | ···················· | ···················· | ···················· | ···················· | ···················· | |
| E | 7897 hCoV-19/England/HECH-259A194/2020\|EPI_ISL_488873\|2 | ···················· | ···················· | ···················· | ···················· | ···················· | ···················· | |
| E | 7898 hCoV-19/Northern Ireland/NIRE-FF1A7/2020\|EPI_ISL_4 | ···················· | ···················· | ···················· | ···················· | ···················· | ···················· | |
| E | 7899 hCoV-19/Northern Ireland/NIRE-FF0B9/2020\|EPI_ISL_4 | ···················· | ···················· | ···················· | ···················· | ···················· | ···················· | |
| E | 7900 hCoV-19/England/BRIS-12DCCC/2020\|EPI_ISL_488877\|20 | ···················· | ···················· | ···················· | ···················· | ···················· | ···················· | |
| E | 7901 hCoV-19/Scotland/EDB3046/2020\|EPI_ISL_488879\|2020- | ···················· | ···················A | ···················· | ···················· | ···················· | ···················· | 1 |
| E | 7902 hCoV-19/Scotland/EDB3128/2020\|EPI_ISL_488880\|2020- | ···················· | ···················· | ···················· | ···················· | ···················· | ···················· | |
| E | 7903 hCoV-19/Scotland/EDB3269/2020\|EPI_ISL_488881\|2020- | ···················· | ···················A | ···················· | ···················· | ···················· | ···················· | 1 |
| E | 7904 hCoV-19/Scotland/EDB3292/2020\|EPI_ISL_488882\|2020- | ···················· | ···················A | ···················· | ···················· | ···················· | ···················· | 1 |
| E | 7905 hCoV-19/Scotland/EDB2956/2020\|EPI_ISL_488883\|2020- | ···················· | ···················A | ···················· | ···················· | ···················· | ···················· | 1 |
| E | 7906 hCoV-19/Scotland/EDB2859/2020\|EPI_ISL_488884\|2020- | ···················· | ···················· | ···················· | ···················· | ···················· | ···················· | |
| E | 7907 hCoV-19/Scotland/EDB3294/2020\|EPI_ISL_488885\|2020- | ···················· | ···················A | ···················· | ···················· | ···················· | ···················· | 1 |
| E | 7908 hCoV-19/Scotland/EDB3159/2020\|EPI_ISL_488890\|2020- | ···················· | ···················· | ···················· | ···················· | ···················· | ···················· | |
| E | 7909 hCoV-19/Scotland/EDB2740/2020\|EPI_ISL_488892\|2020- | ···················· | ···················A | ···················· | ···················· | ···················· | ···················· | 1 |
| E | 7910 hCoV-19/Scotland/EDB3137/2020\|EPI_ISL_488893\|2020- | ···················· | ···················· | ···················· | ···················· | ···················· | ···················· | |
| E | 7911 hCoV-19/Scotland/EDB3223/2020\|EPI_ISL_488894\|2020- | ···················· | ···················· | ···················· | ···················· | ···················· | ···················· | |
| E | 7912 hCoV-19/Scotland/EDB2909/2020\|EPI_ISL_488895\|2020- | ···················· | ···················· | ···················· | ···················· | ···················· | ···················· | |
| E | 7913 hCoV-19/Scotland/EDB3285/2020\|EPI_ISL_488896\|2020- | ···················· | ···················· | ···················· | ···················· | ···················· | ···················· | |
| E | 7914 hCoV-19/Scotland/EDB2915/2020\|EPI_ISL_488897\|2020- | ···················· | ···················· | ···················· | ···················· | ···················· | ···················· | |
| E | 7915 hCoV-19/Scotland/EDB3228/2020\|EPI_ISL_488898\|2020- | ···················· | ···················· | ···················· | ···················· | ···················· | ···················· | |
| E | 7916 hCoV-19/Scotland/EDB2719/2020\|EPI_ISL_488899\|2020- | ···················· | ···················· | ···················· | ···················· | ···················· | ···················· | |
| E | 7917 hCoV-19/USA/WA-S1350/2020\|EPI_ISL_495800\|2020-05-1 | ···················· | ···················· | ···················· | ···················· | ···················· | ···················· | |
| E | 7918 hCoV-19/USA/WA-S1351/2020\|EPI_ISL_495801\|2020-05-1 | ···················· | ···················· | ···················· | ···················· | ···················· | ···················· | |
| E | 7919 hCoV-19/USA/WA-S1352/2020\|EPI_ISL_495802\|2020-05-1 | ···················· | ···················· | ···················· | ···················· | ···················· | ···················· | |
| E | 7920 hCoV-19/USA/WA-S1353/2020\|EPI_ISL_495803\|2020-05-1 | ···················· | ···················· | ···················· | ···················· | ···················· | ···················· | |
| E | 7921 hCoV-19/USA/WA-S1354/2020\|EPI_ISL_495804\|2020-05-1 | ···················· | ···················· | ···················· | ···················· | ···················· | ···················· | |
| E | 7922 hCoV-19/USA/WA-S1355/2020\|EPI_ISL_495805\|2020-05-1 | ···················· | ···················· | ···················· | ···················· | ···················· | ···················· | |
| E | 7923 hCoV-19/USA/WA-S1356/2020\|EPI_ISL_495806\|2020-05-1 | ···················· | ···················· | ···················· | ···················· | ···················· | ···················· | |
| E | 7924 hCoV-19/USA/WA-S1357/2020\|EPI_ISL_495807\|2020-05-1 | ···················· | ···················· | ···················· | ···················· | ···················· | ···················· | |
| E | 7925 hCoV-19/USA/WA-S1358/2020\|EPI_ISL_495808\|2020-05-1 | ···················· | ···················· | ···················· | ···················· | ···················· | ···················· | |
| E | 7926 hCoV-19/USA/WA-S1359/2020\|EPI_ISL_495809\|2020-05-1 | ···················· | ···················· | ···················· | ···················· | ···················· | ···················· | |
| E | 7927 hCoV-19/USA/WA-S1360/2020\|EPI_ISL_495810\|2020-05-1 | ···················· | ···················· | ···················· | ···················· | ···················· | ···················· | |
| E | 7928 hCoV-19/USA/WA-S1361/2020\|EPI_ISL_495811\|2020-05-1 | ···················· | ···················· | ···················· | ···················· | ···················· | ···················· | |
| E | 7929 hCoV-19/USA/WA-S1362/2020\|EPI_ISL_495812\|2020-05-1 | ···················· | ···················· | ···················· | ···················· | ···················· | ···················· | |
| E | 7930 hCoV-19/USA/WA-S1363/2020\|EPI_ISL_495813\|2020-05-1 | ···················· | ···················· | ···················· | ···················· | ···················· | ···················· | |
| E | 7931 hCoV-19/USA/WA-S1364/2020\|EPI_ISL_495814\|2020-05-1 | ···················· | ···················· | ···················· | ···················· | ···················· | ···················· | |
| E | 7932 hCoV-19/USA/WA-S1365/2020\|EPI_ISL_495815\|2020-05-1 | ···················· | ···················· | ···················· | ···················· | ···················· | ···················· | |
| E | 7933 hCoV-19/USA/WA-S1366/2020\|EPI_ISL_495816\|2020-05-1 | ···················· | ···················· | ···················· | ···················· | ···················· | ···················· | |
| E | 7934 hCoV-19/USA/WA-S1367/2020\|EPI_ISL_495817\|2020-05-1 | ···················· | ···················· | ···················· | ···················· | ···················· | ···················· | |
| E | 7935 hCoV-19/USA/WA-S1368/2020\|EPI_ISL_495818\|2020-05-1 | ···················· | ···················· | ···················· | ···················· | ···················· | ···················· | |
| E | 7936 hCoV-19/USA/WA-S1369/2020\|EPI_ISL_495819\|2020-05-1 | ···················· | ···················· | ···················· | ···················· | ···················· | ···················· | |
| E | 7937 hCoV-19/Japan/PG-0337/2020\|EPI_ISL_480010\|2020-04- | ···················· | ···················· | ···················· | ···················· | ···················· | ···················· | |
| E | 7938 hCoV-19/Japan/PG-0338/2020\|EPI_ISL_480011\|2020-03- | ···················· | ···················· | ···················· | ···················· | ···················· | ···················· | |
| E | 7939 hCoV-19/Japan/PG-0339/2020\|EPI_ISL_480012\|2020-03- | ···················· | ···················· | ···················· | ···················· | ···················· | ···················· | |
| E | 7940 hCoV-19/Japan/PG-0340/2020\|EPI_ISL_480013\|2020-03- | ···················· | ···················· | ···················· | ···················· | ···················· | ···················· | |
| E | 7941 hCoV-19/Japan/PG-0342/2020\|EPI_ISL_480014\|2020-03- | ···················· | ···················· | ···················· | ···················· | ···················· | ···················· | |
| E | 7942 hCoV-19/Japan/PG-0343/2020\|EPI_ISL_480015\|2020-03- | ···················· | ···················· | ···················· | ···················· | ···················· | ···················· | |
| E | 7943 hCoV-19/Japan/PG-0344/2020\|EPI_ISL_480016\|2020-03- | ···················· | ···················· | ···················· | ···················· | ···················· | ···················· | |
| E | 7944 hCoV-19/Japan/PG-0345/2020\|EPI_ISL_480017\|2020-03- | ···················· | ···················· | ···················· | ···················· | ···················· | ···················· | |
| E | 7945 hCoV-19/Japan/PG-0346/2020\|EPI_ISL_480018\|2020-03- | ···················· | ···················· | ···················· | ···················· | ···················· | ···················· | |
| E | 7946 hCoV-19/Japan/PG-0347/2020\|EPI_ISL_480019\|2020-03- | ···················· | ···················· | ···················· | ···················· | ···················· | ···················· | |
| E | 7947 hCoV-19/Japan/PG-0348/2020\|EPI_ISL_480020\|2020-03- | ···················· | ···················· | ···················· | ···················· | ···················· | ···················· | |
| E | 7948 hCoV-19/Japan/PG-0349/2020\|EPI_ISL_480021\|2020-03- | ···················· | ···················· | ···················· | ···················· | ···················· | ···················· | |
| E | 7949 hCoV-19/Japan/PG-0350/2020\|EPI_ISL_480022\|2020-03- | ···················· | ···················· | ···················· | ···················· | ···················· | ···················· | |
| E | 7950 hCoV-19/Japan/PG-0351/2020\|EPI_ISL_480023\|2020-03- | ···················· | ···················· | ···················· | ···················· | ···················· | ···················· | |
| E | 7951 hCoV-19/Japan/PG-0352/2020\|EPI_ISL_480024\|2020-03- | ···················· | ···················· | ···················· | ···················· | ···················· | ···················· | |
| E | 7952 hCoV-19/Japan/PG-0353/2020\|EPI_ISL_480025\|2020-03- | ···················· | ···················· | ···················· | ···················· | ···················· | ···················· | |
| E | 7953 hCoV-19/Japan/PG-0354/2020\|EPI_ISL_480026\|2020-03- | ···················· | ···················· | ···················· | ···················· | ···················· | ···················· | |
| E | 7954 hCoV-19/Japan/PG-0355/2020\|EPI_ISL_480027\|2020-03- | ···················· | ···················· | ···················· | ···················· | ···················· | ···················· | |
| E | 7955 hCoV-19/Japan/PG-0356/2020\|EPI_ISL_480028\|2020-03- | ···················· | ···················· | ···················· | ···················· | ···················· | ···················· | |
| E | 7956 hCoV-19/Japan/PG-0357/2020\|EPI_ISL_480029\|2020-04- | ···················· | ···················· | ···················· | ···················· | ···················· | ···················· | |
| E | 7957 hCoV-19/Japan/PG-0359/2020\|EPI_ISL_480030\|2020-02- | ···················· | ···················· | ···················· | ···················· | ···················· | ···················· | |

| | | | | | | | | |
|---|---|---|---|---|---|---|---|---|
| E | 7958 | hCoV-19/Japan/PG-0384/2020\|EPI_ISL_480053\|2020-03- | ···················· | ···················· | ····················· | ···················· | ···················· | ···················· |
| E | 7959 | hCoV-19/Japan/PG-0385/2020\|EPI_ISL_480054\|2020-03- | ···················· | ···················· | ····················· | ···················· | ···················· | ···················· |
| E | 7960 | hCoV-19/Japan/PG-0386/2020\|EPI_ISL_480055\|2020-03- | ···················· | ···················· | ····················· | ···················· | ···················· | ···················· |
| E | 7961 | hCoV-19/Japan/PG-0387/2020\|EPI_ISL_480056\|2020-03- | ···················· | ···················· | ····················· | ···················· | ···················· | ···················· |
| E | 7962 | hCoV-19/Japan/PG-0388/2020\|EPI_ISL_480057\|2020-03- | ···················· | ···················· | ····················· | ···················· | ···················· | ···················· |
| E | 7963 | hCoV-19/Japan/PG-0389/2020\|EPI_ISL_480058\|2020-03- | ···················· | ···················· | ····················· | ···················· | ···················· | ···················· |
| E | 7964 | hCoV-19/Japan/PG-0390/2020\|EPI_ISL_480059\|2020-03- | ···················· | ···················· | ····················· | ···················· | ···················· | ···················· |
| E | 7965 | hCoV-19/Japan/PG-0391/2020\|EPI_ISL_480060\|2020-03- | ···················· | ···················· | ····················· | ···················· | ···················· | ···················· |
| E | 7966 | hCoV-19/Japan/PG-0392/2020\|EPI_ISL_480061\|2020-03- | ···················· | ···················· | ····················· | ···················· | ···················· | ···················· |
| E | 7967 | hCoV-19/Japan/PG-0393/2020\|EPI_ISL_480062\|2020-03- | ···················· | ···················· | ····················· | ···················· | ···················· | ···················· |
| E | 7968 | hCoV-19/Japan/PG-0394/2020\|EPI_ISL_480063\|2020-03- | ···················· | ···················· | ····················· | ···················· | ···················· | ···················· |
| E | 7969 | hCoV-19/Japan/PG-0395/2020\|EPI_ISL_480064\|2020-03- | ···················· | ···················· | ····················· | ···················· | ···················· | ···················· |
| E | 7970 | hCoV-19/Japan/PG-0396/2020\|EPI_ISL_480065\|2020-03- | ···················· | ···················· | ····················· | ···················· | ···················· | ···················· |
| E | 7971 | hCoV-19/Japan/PG-0397/2020\|EPI_ISL_480066\|2020-03- | ···················· | ···················· | ····················· | ···················· | ···················· | ···················· |
| E | 7972 | hCoV-19/Japan/PG-0398/2020\|EPI_ISL_480067\|2020-03- | ···················· | ···················· | ····················· | ···················· | ···················· | ···················· |
| E | 7973 | hCoV-19/Japan/PG-0399/2020\|EPI_ISL_480068\|2020-04- | ···················· | ···················· | ····················· | ···················· | ···················· | ···················· |
| E | 7974 | hCoV-19/Japan/PG-0400/2020\|EPI_ISL_480069\|2020-04- | ···················· | ···················· | ····················· | ···················· | ···················· | ···················· |
| E | 7975 | hCoV-19/Japan/PG-0401/2020\|EPI_ISL_480070\|2020-04- | ···················· | ···················· | ····················· | ···················· | ···················· | ···················· |
| E | 7976 | hCoV-19/Japan/PG-0402/2020\|EPI_ISL_480071\|2020-04- | ···················· | ···················· | ····················· | ···················· | ···················· | ···················· |
| E | 7977 | hCoV-19/Japan/PG-0403/2020\|EPI_ISL_480072\|2020-04- | ···················· | ···················· | ····················· | ···················· | ···················· | ···················· |
| E | 7978 | hCoV-19/Japan/PG-0404/2020\|EPI_ISL_480073\|2020-03- | ···················· | ···················· | ····················· | ···················· | ···················· | ···················· |
| E | 7979 | hCoV-19/Japan/PG-0405/2020\|EPI_ISL_480074\|2020-03- | ···················· | ···················· | ····················· | ···················· | ···················· | ···················· |
| E | 7980 | hCoV-19/Japan/PG-0406/2020\|EPI_ISL_480075\|2020-03- | ···················· | ···················· | ····················· | ···················· | ···················· | ···················· |
| E | 7981 | hCoV-19/Japan/PG-0407/2020\|EPI_ISL_480076\|2020-03- | ···················· | ···················· | ····················· | ···················· | ···················· | ···················· |
| E | 7982 | hCoV-19/Japan/PG-0408/2020\|EPI_ISL_480077\|2020-03- | ···················· | ···················· | ····················· | ···················· | ···················· | ···················· |
| E | 7983 | hCoV-19/Japan/PG-0409/2020\|EPI_ISL_480078\|2020-03- | ···················· | ···················· | ····················· | ···················· | ···················· | ···················· |
| E | 7984 | hCoV-19/Japan/PG-0410/2020\|EPI_ISL_480079\|2020-03- | ···················· | ···················· | ····················· | ···················· | ···················· | ···················· |
| E | 7985 | hCoV-19/Japan/PG-0411/2020\|EPI_ISL_480080\|2020-03- | ···················· | ···················· | ····················· | ···················· | ···················· | ···················· |
| E | 7986 | hCoV-19/Japan/PG-0412/2020\|EPI_ISL_480081\|2020-03- | ···················· | ···················· | ····················· | ···················· | ···················· | ···················· |
| E | 7987 | hCoV-19/Japan/PG-0413/2020\|EPI_ISL_480082\|2020-04- | ···················· | ···················· | ····················· | ···················· | ···················· | ···················· |
| E | 7988 | hCoV-19/Japan/PG-0415/2020\|EPI_ISL_480083\|2020-03- | ···················· | ···················· | ····················· | ···················· | ···················· | ···················· |
| E | 7989 | hCoV-19/Japan/PG-0416/2020\|EPI_ISL_480084\|2020-03- | ···················· | ···················· | ····················· | ···················· | ···················· | ···················· |
| E | 7990 | hCoV-19/Japan/PG-0417/2020\|EPI_ISL_480085\|2020-03- | ···················· | ···················· | ····················· | ···················· | ···················· | ···················· |
| E | 7991 | hCoV-19/Japan/PG-0418/2020\|EPI_ISL_480086\|2020-03- | ···················· | ···················· | ····················· | ···················· | ···················· | ···················· |
| E | 7992 | hCoV-19/Japan/PG-0419/2020\|EPI_ISL_480087\|2020-03- | ···················· | ···················· | ····················· | ···················· | ···················· | ···················· |
| E | 7993 | hCoV-19/Japan/PG-0420/2020\|EPI_ISL_480088\|2020-03- | ···················· | ···················· | ····················· | ···················· | ···················· | ···················· |
| E | 7994 | hCoV-19/Japan/PG-0421/2020\|EPI_ISL_480089\|2020-03- | ···················· | ···················· | ····················· | ···················· | ···················· | ···················· |
| E | 7995 | hCoV-19/Japan/PG-0422/2020\|EPI_ISL_480090\|2020-04- | ···················· | ···················· | ····················· | ···················· | ···················· | ···················· |
| E | 7996 | hCoV-19/Japan/PG-0423/2020\|EPI_ISL_480091\|2020-04- | ···················· | ···················· | ····················· | ···················· | ···················· | ···················· |
| E | 7997 | hCoV-19/Japan/PG-0424/2020\|EPI_ISL_480092\|2020-04- | ···················· | ···················· | ····················· | ···················· | ···················· | ···················· |
| E | 7998 | hCoV-19/Japan/PG-0425/2020\|EPI_ISL_480093\|2020-04- | ···················· | ···················· | ····················· | ···················· | ···················· | ···················· |
| E | 7999 | hCoV-19/Japan/PG-0426/2020\|EPI_ISL_480094\|2020-04- | ···················· | ···················· | ····················· | ···················· | ···················· | ···················· |
| E | 8000 | hCoV-19/Japan/PG-0427/2020\|EPI_ISL_480095\|2020-04- | ···················· | ···················· | ····················· | ···················· | ···················· | ···················· |
| E | 8001 | hCoV-19/Japan/PG-0430/2020\|EPI_ISL_480097\|2020-04- | ···················· | ···················· | ····················· | ···················· | ···················· | ···················· |
| E | 8002 | hCoV-19/Japan/PG-0432/2020\|EPI_ISL_480098\|2020-04- | ···················· | ···················· | ····················· | ···················· | ···················· | ···················· |
| E | 8003 | hCoV-19/Japan/PG-0433/2020\|EPI_ISL_480099\|2020-04- | ···················· | ···················· | ····················· | ···················· | ···················· | ···················· |
| E | 8004 | hCoV-19/Singapore/24nan/2020\|EPI_ISL_482672\|2020-0 | ···················· | ···················· | ····················· | ···················· | ···················· | ···················· |
| E | 8005 | hCoV-19/Singapore/372/2020\|EPI_ISL_482673\|2020-04- | ···················· | ···················· | ····················· | ···················· | ···················· | ···················· |
| E | 8006 | hCoV-19/Singapore/371/2020\|EPI_ISL_482674\|2020-04- | ···················· | ···················· | ····················· | ···················· | ···················· | ···················· |
| E | 8007 | hCoV-19/Singapore/370/2020\|EPI_ISL_482675\|2020-04- | ···················· | ···················· | ····················· | ···················· | ···················· | ···················· |
| E | 8008 | hCoV-19/Singapore/368nan/2020\|EPI_ISL_482676\|2020- | ···················· | ···················· | ····················· | ···················· | ···················· | ···················· |
| E | 8009 | hCoV-19/Singapore/367/2020\|EPI_ISL_482677\|2020-03- | ···················· | ···················· | ····················· | ···················· | ···················· | ···················· |
| E | 8011 | hCoV-19/Singapore/544/2020\|EPI_ISL_482682\|2020-06- | ···················· | ···················· | ····················· | ···················· | ···················· | ···················· |
| E | 8016 | hCoV-19/Singapore/539/2020\|EPI_ISL_482687\|2020-06- | ···················· | ···················· | ····················· | ···················· | ···················· | N······N············ |
| E | 8021 | hCoV-19/Singapore/545/2020\|EPI_ISL_482692\|2020-05- | ···················· | A··················· | ····················· | ···················· | ···················· | ···················· |
| E | 8022 | hCoV-19/USA/CA-CZB-1946/2020\|EPI_ISL_486224\|2020-0 | ···················· | ···················· | ····················· | ···················· | ···················· | ···················· |
| E | 8023 | hCoV-19/USA/CA-CZB-1949/2020\|EPI_ISL_486226\|2020-0 | ···················· | ···················· | ····················· | ···················· | ···················· | ···················· |
| E | 8024 | hCoV-19/USA/CA-CZB-1955/2020\|EPI_ISL_486228\|2020-0 | ···················· | ···················· | ····················· | ···················· | ···················· | ···················· |
| E | 8025 | hCoV-19/USA/CA-CZB-1957/2020\|EPI_ISL_486229\|2020-0 | ···················· | ···················· | ····················· | ···················· | ···················· | ···················· |
| E | 8026 | hCoV-19/USA/CA-CZB-1958/2020\|EPI_ISL_486230\|2020-0 | ···················· | ···················· | ····················· | ···················· | ···················· | ···················· |
| E | 8027 | hCoV-19/England/BRIS-12B3E7/2020\|EPI_ISL_487555\|20 | ···················· | ···················· | ····················· | ···················· | ···················· | ···················· |
| E | 8028 | hCoV-19/England/LOND-D66A7/2020\|EPI_ISL_487558\|202 | ···················· | ·········A·········· | ····················· | ···················· | ···················· | ···················· |
| E | 8029 | hCoV-19/England/LOND-D6425/2020\|EPI_ISL_487559\|202 | ···················· | ···················· | ····················· | ···················· | ···················· | ···················· |
| E | 8030 | hCoV-19/England/BRIS-12B341/2020\|EPI_ISL_487560\|20 | ···················· | ···················· | ····················· | ···················· | ···················· | ···················· |
| E | 8031 | hCoV-19/England/NORT-281041/2020\|EPI_ISL_488800\|20 | ···················· | ···················· | ····················· | ···················· | ···················· | ···················· |
| E | 8032 | hCoV-19/England/NORT-281524/2020\|EPI_ISL_488801\|20 | ···················· | ···················· | ····················· | ···················· | ···················· | ···················· |
| E | 8033 | hCoV-19/England/NORT-281C98/2020\|EPI_ISL_488802\|20 | ···················· | ···················· | ····················· | ···················· | ···················· | ···················· |
| E | 8034 | hCoV-19/England/NORT-28106F/2020\|EPI_ISL_488803\|20 | ···················· | ···················· | ····················· | ···················· | ···················· | ···················· |
| E | 8035 | hCoV-19/England/NORT-281849/2020\|EPI_ISL_488804\|20 | ···················· | ···················· | ····················· | ···················· | ···················· | ···················· |
| E | 8036 | hCoV-19/England/NORT-281630/2020\|EPI_ISL_488805\|20 | ···················· | ···················· | ····················· | ···················· | ···················· | ···················· |
| E | 8037 | hCoV-19/England/NORT-2813A2/2020\|EPI_ISL_488806\|20 | ···················· | ·············A······ | ····················· | ···················· | ···················· | ···················· |
| E | 8038 | hCoV-19/England/NORT-28124B/2020\|EPI_ISL_488807\|20 | ···················· | ···················· | ····················· | ···················· | ···················· | ···················· |
| E | 8039 | hCoV-19/England/NORT-283E90/2020\|EPI_ISL_488808\|20 | ···················· | ···················· | ····················· | ···················· | ···················· | ···················· |
| E | 8040 | hCoV-19/England/NORT-281603/2020\|EPI_ISL_488809\|20 | ···················· | ···················· | ····················· | ···················· | ···················· | ···················· |
| E | 8041 | hCoV-19/England/NORT-281885/2020\|EPI_ISL_488810\|20 | ···················· | ···················· | ····················· | ···················· | ···················· | ···················· |
| E | 8042 | hCoV-19/England/NORT-281894/2020\|EPI_ISL_488811\|20 | ···················· | ···················· | ····················· | ···················· | ···················· | ···················· |
| E | 8043 | hCoV-19/England/NORT-281946/2020\|EPI_ISL_488812\|20 | ···················· | ···················· | ····················· | ···················· | ···················· | ···················· |
| E | 8044 | hCoV-19/England/NORT-2819FB/2020\|EPI_ISL_488813\|20 | ···················· | ···················· | ····················· | ···················· | ···················· | ···················· |
| E | 8045 | hCoV-19/England/NORT-2818D0/2020\|EPI_ISL_488814\|20 | ···················· | ···················· | ····················· | ···················· | ···················· | ···················· |
| E | 8046 | hCoV-19/England/NORT-2819DD/2020\|EPI_ISL_488815\|20 | ···················· | ···················· | ····················· | ···················· | ···················· | ···················· |

| | | | | | | | | |
|---|---|---|---|---|---|---|---|---|
| E | 8047 hCoV-19/England/NORT-281ACB/2020\|EPI_ISL_488816\|20 | .................... | .................... | .................... | .................... | .................... | .................... | |
| E | 8048 hCoV-19/England/NORT-281120/2020\|EPI_ISL_488817\|20 | .................... | .................... | .................... | .................... | .................... | .................... | |
| E | 8049 hCoV-19/England/NORT-281797/2020\|EPI_ISL_488818\|20 | .................... | .................... | .................... | .................... | .................... | .................... | |
| E | 8050 hCoV-19/England/NORT-281C01/2020\|EPI_ISL_488819\|20 | .................... | .A.................. | .................... | .................... | .................... | .................... | |
| E | 8051 hCoV-19/England/NORT-2841A5/2020\|EPI_ISL_488820\|20 | .................... | .................... | .................... | .................... | .................... | .................... | |
| E | 8052 hCoV-19/England/NORT-2817D3/2020\|EPI_ISL_488821\|20 | .................... | .................... | .................... | .................... | .................... | .................... | |
| E | 8053 hCoV-19/England/NORT-281A9E/2020\|EPI_ISL_488822\|20 | .................... | .................... | .................... | .................... | .................... | .................... | |
| E | 8054 hCoV-19/England/NORT-28168B/2020\|EPI_ISL_488823\|20 | .................... | .................... | .................... | .................... | .................... | .................... | |
| E | 8055 hCoV-19/England/NORT-284354/2020\|EPI_ISL_488824\|20 | .................... | .................... | .................... | .................... | .................... | .................... | |
| E | 8056 hCoV-19/England/NORT-2819A0/2020\|EPI_ISL_488825\|20 | .................... | .................... | .................... | .................... | .................... | .................... | |
| E | 8057 hCoV-19/England/NORT-281418/2020\|EPI_ISL_488826\|20 | .................... | .................... | .................... | .................... | .................... | .................... | |
| E | 8058 hCoV-19/England/NORT-283BE4/2020\|EPI_ISL_488827\|20 | .................... | .................... | .................... | .................... | .................... | .................... | |
| E | 8059 hCoV-19/England/NORT-28164F/2020\|EPI_ISL_488828\|20 | .................... | .................... | .................... | .................... | .................... | .................... | |
| E | 8060 hCoV-19/England/NORT-2811C6/2020\|EPI_ISL_488829\|20 | .................... | .................... | .................... | .................... | .................... | .................... | |
| E | 8061 hCoV-19/England/NORT-281B40/2020\|EPI_ISL_488830\|20 | .................... | .................... | .................... | .................... | .................... | .................... | |
| E | 8062 hCoV-19/England/NORT-28115D/2020\|EPI_ISL_488831\|20 | .................... | .................... | .................... | .................... | .................... | .................... | |
| E | 8063 hCoV-19/England/NORT-281973/2020\|EPI_ISL_488832\|20 | .................... | .................... | .................... | .................... | .................... | .................... | |
| E | 8064 hCoV-19/England/NORT-281384/2020\|EPI_ISL_488834\|20 | .................... | .................... | .................... | .................... | .................... | .................... | |
| E | 8065 hCoV-19/England/NORT-283C2D/2020\|EPI_ISL_488835\|20 | .................... | .................... | .................... | .................... | .................... | .................... | |
| E | 8066 hCoV-19/England/NORT-283DB1/2020\|EPI_ISL_488836\|20 | .................... | .................... | .................... | .................... | .................... | .................... | |
| E | 8067 hCoV-19/England/HECH-259A11C/2020\|EPI_ISL_488837\|2 | .................... | .................... | .................... | .................... | .................... | .................... | |
| E | 8068 hCoV-19/England/HECH-259A1D0/2020\|EPI_ISL_488838\|2 | .................... | .................... | .................... | .................... | .................... | .................... | |
| E | 8069 hCoV-19/England/HECH-259A158/2020\|EPI_ISL_488839\|2 | .................... | .................... | .................... | .................... | .................... | .................... | |
| E | 8070 hCoV-19/England/WSFT-25C19D8/2020\|EPI_ISL_488840\|2 | .................... | .................... | .................... | .................... | .................... | .................... | |
| E | 8071 hCoV-19/England/HECH-259A10D/2020\|EPI_ISL_488842\|2 | .................... | .................... | .................... | .................... | .................... | .................... | |
| E | 8072 hCoV-19/England/HECH-259A0E2/2020\|EPI_ISL_488843\|2 | .................... | .................... | .................... | .................... | .................... | .................... | |
| E | 8073 hCoV-19/England/HECH-259A273/2020\|EPI_ISL_488844\|2 | .................... | .................... | .................... | .................... | .................... | .................... | |
| E | 8074 hCoV-19/England/WSFT-25C19AB/2020\|EPI_ISL_488845\|2 | .................... | .................... | .................... | .................... | .................... | .................... | |
| E | 8075 hCoV-19/England/HECH-259A3DA/2020\|EPI_ISL_488848\|2 | .................... | .................... | .................... | .................... | .................... | .................... | |
| E | 8076 hCoV-19/England/HECH-259A3E9/2020\|EPI_ISL_488850\|2 | .................... | .................... | .................... | .................... | .................... | .................... | |
| E | 8077 hCoV-19/England/HECH-259A0B5/2020\|EPI_ISL_488851\|2 | .................... | .................... | .................... | ......C............. | .................... | .................... | |
| E | 8078 hCoV-19/England/HECH-259A0D3/2020\|EPI_ISL_488852\|2 | .................... | .................... | .................... | ......C............. | .................... | .................... | |
| E | 8079 hCoV-19/Scotland/EDB2917/2020\|EPI_ISL_488886\|2020- | .................... | ...................A | .................... | .................... | .................... | .................... | 1 |
| E | 8080 hCoV-19/Scotland/EDB2797/2020\|EPI_ISL_488887\|2020- | .................... | .................... | .................... | .................... | .................... | .................... | |
| E | 8081 hCoV-19/Scotland/EDB3077/2020\|EPI_ISL_488888\|2020- | .................... | .................... | .................... | .................... | .................... | .................... | |
| E | 8082 hCoV-19/Scotland/EDB3033/2020\|EPI_ISL_488889\|2020- | .................... | .................... | .................... | .................... | .................... | .................... | |
| E | 8083 hCoV-19/Scotland/EDB3023/2020\|EPI_ISL_488891\|2020- | .................... | .................... | .................... | .................... | .................... | .................... | |
| E | 8084 hCoV-19/Oman/205038896/2020\|EPI_ISL_492000\|2020-05 | .................... | .................... | .................... | .................... | .................... | .................... | |
| E | 8085 hCoV-19/Oman/205038906/2020\|EPI_ISL_492001\|2020-05 | .................... | .................... | .................... | .................... | .................... | .................... | |
| E | 8086 hCoV-19/Oman/205039048/2020\|EPI_ISL_492002\|2020-05 | .................... | .................... | .................... | .................... | .................... | .................... | |
| E | 8087 hCoV-19/Oman/205039149/2020\|EPI_ISL_492003\|2020-05 | .................... | .................... | .................... | .................... | .................... | .................... | |
| E | 8088 hCoV-19/Oman/205039531/2020\|EPI_ISL_492005\|2020-05 | .................... | .................... | .................... | .................... | .................... | .................... | |
| E | 8089 hCoV-19/Oman/205039569/2020\|EPI_ISL_492006\|2020-05 | .................... | .................... | .................... | .................... | .................... | .................... | |
| E | 8090 hCoV-19/Oman/205039772/2020\|EPI_ISL_492007\|2020-05 | .................... | .................... | .................... | .................... | .................... | .................... | |
| E | 8091 hCoV-19/Oman/205039788/2020\|EPI_ISL_492008\|2020-05 | .................... | .................... | .................... | .................... | .................... | .................... | |
| E | 8092 hCoV-19/Oman/205039804/2020\|EPI_ISL_492009\|2020-05 | .................... | .................... | .................... | .................... | .................... | .................... | |
| E | 8093 hCoV-19/Brazil/RJ-UFRJ-1019/2020\|EPI_ISL_492032\|20 | .................... | .................... | .................... | .................... | .................... | .................... | |
| E | 8094 hCoV-19/Brazil/RJ-UFRJ-1640/2020\|EPI_ISL_492033\|20 | .................... | .................... | .................... | .................... | .................... | .................... | |
| E | 8095 hCoV-19/Brazil/RJ-UFRJ-2215/2020\|EPI_ISL_492034\|20 | .................... | .................... | .................... | .................... | .................... | .................... | |
| E | 8096 hCoV-19/Brazil/RJ-UFRJ-2517/2020\|EPI_ISL_492035\|20 | .................... | .................... | .................... | .................... | .................... | .................... | |
| E | 8097 hCoV-19/Brazil/RJ-UFRJ-9331/2020\|EPI_ISL_492036\|20 | .................... | .................... | .................... | .................... | .................... | .................... | |
| E | 8098 hCoV-19/Brazil/RJ-UFRJ-1784/2020\|EPI_ISL_492037\|20 | .................... | .................... | .................... | .................... | .................... | .................... | |
| E | 8099 hCoV-19/Brazil/RJ-UFRJ-11785/2020\|EPI_ISL_492038\|2 | .................... | .................... | .................... | .................... | .................... | .................... | |
| E | 8100 hCoV-19/Brazil/RJ-UFRJ-11786/2020\|EPI_ISL_492039\|2 | .................... | .................... | .................... | .................... | .................... | .................... | |
| E | 8101 hCoV-19/Brazil/RJ-UFRJ-32317/2020\|EPI_ISL_492040\|2 | .................... | .................... | .................... | .................... | .................... | .................... | |
| E | 8102 hCoV-19/Brazil/RJ-UFRJ-32321/2020\|EPI_ISL_492041\|2 | .................... | .................... | .................... | .................... | .................... | .................... | |
| E | 8103 hCoV-19/Brazil/RJ-UFRJ-54093/2020\|EPI_ISL_492042\|2 | .................... | .................... | .................... | .................... | .................... | .................... | |
| E | 8104 hCoV-19/Brazil/RJ-UFRJ-57437/2020\|EPI_ISL_492043\|2 | .................... | .................... | .................... | .................... | .................... | .................... | |
| E | 8105 hCoV-19/Brazil/RJ-UFRJ-57721/2020\|EPI_ISL_492044\|2 | .................... | .................... | .................... | .................... | .................... | .................... | |
| E | 8106 hCoV-19/Brazil/RJ-UFRJ-57722/2020\|EPI_ISL_492045\|2 | .................... | .................... | .................... | .................... | .................... | .................... | |
| E | 8107 hCoV-19/Brazil/RJ-UFRJ-58090/2020\|EPI_ISL_492046\|2 | .................... | .................... | .................... | .................... | .................... | .................... | |
| E | 8108 hCoV-19/Brazil/RJ-UFRJ-58110/2020\|EPI_ISL_492047\|2 | .................... | .................... | .................... | .................... | .................... | .................... | |
| E | 8109 hCoV-19/Brazil/RJ-UFRJ-58271/2020\|EPI_ISL_492048\|2 | .................... | .................... | .................... | .................... | .................... | .................... | |
| E | 8110 hCoV-19/USA/AK-PHL108/2020\|EPI_ISL_492059\|2020-04- | .................... | .................... | .................... | .................... | .................... | .................... | |
| E | 8111 hCoV-19/USA/AK-PHL280/2020\|EPI_ISL_492063\|2020-04- | .................... | .................... | .................... | .................... | .................... | .................... | |
| E | 8112 hCoV-19/Oman/810962/2020\|EPI_ISL_492065\|2020-06-20 | .................... | .................... | .................... | .................... | .................... | .................... | |
| E | 8113 hCoV-19/Poland/PL_P41/2020\|EPI_ISL_492066\|2020-04- | .................... | .................... | .................... | .................... | .................... | .................... | |
| E | 8114 hCoV-19/Poland/PL_P42/2020\|EPI_ISL_492067\|2020-03- | .................... | .................... | .................... | .................... | .................... | .................... | |
| E | 8115 hCoV-19/Poland/PL_P43/2020\|EPI_ISL_492068\|2020-04- | .................... | .................... | .................... | .................... | .................... | .................... | |
| E | 8116 hCoV-19/Poland/PL_P44/2020\|EPI_ISL_492069\|2020-04- | .................... | .................... | .................... | .................... | .................... | .................... | |
| E | 8117 hCoV-19/Poland/PL_P45/2020\|EPI_ISL_492070\|2020-04- | .................... | .................... | .................... | .................... | .................... | .................... | |
| E | 8118 hCoV-19/Poland/PL_P46/2020\|EPI_ISL_492071\|2020-04- | .................... | .................... | .................... | .................... | .................... | .................... | |
| E | 8119 hCoV-19/Poland/PL_P47/2020\|EPI_ISL_492072\|2020-04- | .................... | .................... | .................... | .................... | .................... | .................... | |
| E | 8120 hCoV-19/Poland/PL_P48/2020\|EPI_ISL_492073\|2020-04- | .................... | .................... | .................... | .................... | .................... | .................... | |
| E | 8121 hCoV-19/USA/SC-USC-PH00769/2020\|EPI_ISL_492074\|202 | .................... | .........N.......... | .................... | .................... | .................... | .................... | |
| E | 8122 hCoV-19/North Macedonia/ChVir7903/2020\|EPI_ISL_492 | .................... | .................... | .................... | .................... | .................... | .................... | |
| E | 8123 hCoV-19/North Macedonia/ChVir7904/2020\|EPI_ISL_492 | .................... | ....G............... | .................... | .................... | .................... | .................... | |
| E | 8124 hCoV-19/North Macedonia/ChVir7905/2020\|EPI_ISL_492 | .................... | .................... | .................... | .................... | .................... | .................... | |
| E | 8125 hCoV-19/North Macedonia/ChVir7907/2020\|EPI_ISL_492 | .................... | .................... | .................... | .................... | .................... | .................... | |
| E | 8126 hCoV-19/North Macedonia/ChVir7908/2020\|EPI_ISL_492 | .................... | .................... | .................... | .................... | .................... | .................... | |

| | | | | | | | | |
|---|---|---|---|---|---|---|---|---|
| E | 8127 hCoV-19/North Macedonia/ChVir7909/2020\|EPI_ISL_492 | ···················· | ···················· | ···················· | ···················· | ···················· | ···················· | |
| E | 8128 hCoV-19/North Macedonia/ChVir7911/2020\|EPI_ISL_492 | ···················· | ···················· | ···················· | ···················· | ···················· | ···················· | |
| E | 8129 hCoV-19/North Macedonia/ChVir7912/2020\|EPI_ISL_492 | ···················· | ···················· | ···················· | ···················· | ···················· | ···················· | |
| E | 8130 hCoV-19/North Macedonia/ChVir7913/2020\|EPI_ISL_492 | ···················· | ···················· | ···················· | ···················· | ···················· | ···················· | |
| E | 8131 hCoV-19/North Macedonia/ChVir7914/2020\|EPI_ISL_492 | ···················· | ···················· | ···················· | ···················· | ···················· | ···················· | |
| E | 8132 hCoV-19/North Macedonia/ChVir7915/2020\|EPI_ISL_492 | ···················· | ···················· | ···················· | ···················· | ···················· | ···················· | |
| E | 8133 hCoV-19/North Macedonia/ChVir7918/2020\|EPI_ISL_492 | ···················· | ···················· | ···················· | ···················· | ···················· | ···················· | |
| E | 8134 hCoV-19/Italy/LAZ-INMI16/2020\|EPI_ISL_493328\|2020- | ···················· | ···················· | ···················· | ···················· | ···················· | ···················· | |
| E | 8135 hCoV-19/Italy/LAZ-INMI17/2020\|EPI_ISL_493329\|2020- | ···················· | ···················· | ···················· | ···················· | ···················· | ···················· | |
| E | 8136 hCoV-19/Italy/LAZ-INMI18/2020\|EPI_ISL_493330\|2020- | ···················· | ···················· | ···················· | ···················· | ···················· | ···················· | |
| E | 8137 hCoV-19/Italy/LAZ-INMI19/2020\|EPI_ISL_493331\|2020- | ···················· | ···················· | ···················· | ···················· | ···················· | ···················· | |
| E | 8138 hCoV-19/Italy/CAM-INMI-32798-45/2020\|EPI_ISL_49333 | ···················· | ···················· | ···················· | ···················· | ···················· | ···················· | |
| E | 8139 hCoV-19/Italy/CAM-INMI-32803-66/2020\|EPI_ISL_49333 | ···················· | ···················· | ···················· | ···················· | ···················· | ···················· | |
| E | 8141 hCoV-19/Mexico/GUA-InDRE-20/2020\|EPI_ISL_493335\|20 | ···················· | ···················· | ···················· | ···················· | ···················· | ···················· | |
| E | 8142 hCoV-19/Mexico/SIN-InDRE-21/2020\|EPI_ISL_493336\|20 | ···················· | ···················· | ···················· | ···················· | ···················· | ···················· | |
| E | 8143 hCoV-19/Norway/2797/2020\|EPI_ISL_493363\|2020-06-26 | ···················· | ·A·················· | ···················· | ···················· | ···················· | ···················· | |
| E | 8144 hCoV-19/Norway/2798/2020\|EPI_ISL_493364\|2020-06-26 | ···················· | ···················· | ···················· | ···················· | ···················· | ···················· | |
| E | 8145 hCoV-19/Norway/2799/2020\|EPI_ISL_493365\|2020-06-26 | ···················· | ·A·················· | ···················· | ···················· | ···················· | ···················· | |
| E | 8146 hCoV-19/Norway/2800/2020\|EPI_ISL_493366\|2020-06-26 | ···················· | ···················· | ···················· | ···················· | ···················· | ···················· | |
| E | 8147 hCoV-19/Norway/2801/2020\|EPI_ISL_493367\|2020-06-26 | ···················· | ·A·················· | ···················· | ···················· | ···················· | ···················· | |
| E | 8148 hCoV-19/Norway/2802/2020\|EPI_ISL_493368\|2020-06-26 | ···················· | ·A·················· | ···················· | ···················· | ···················· | ···················· | |
| E | 8149 hCoV-19/Norway/2805/2020\|EPI_ISL_493370\|2020-06-27 | ···················· | ·A·················· | ···················· | ···················· | ···················· | ···················· | |
| E | 8150 hCoV-19/Norway/2806/2020\|EPI_ISL_493371\|2020-06-26 | ···················· | ·A·················· | ···················· | ···················· | ···················· | ···················· | |
| E | 8151 hCoV-19/Norway/2807/2020\|EPI_ISL_493372\|2020-06-29 | ···················· | ·A·················· | ···················· | ···················· | ···················· | ···················· | |
| E | 8152 hCoV-19/Norway/2808/2020\|EPI_ISL_493373\|2020-06-29 | ···················· | ·A·················· | ···················· | ···················· | ···················· | ···················· | |
| E | 8153 hCoV-19/Norway/2810/2020\|EPI_ISL_493375\|2020-06-29 | ···················· | ·A·················· | ···················· | ···················· | ···················· | ···················· | |
| E | 8154 hCoV-19/Norway/2811/2020\|EPI_ISL_493376\|2020-06-29 | ···················· | ·A·················· | ···················· | ···················· | ···················· | ···················· | |
| E | 8155 hCoV-19/Norway/2812/2020\|EPI_ISL_493377\|2020-06-29 | ···················· | ·A·················· | ···················· | ···················· | ···················· | ···················· | |
| E | 8156 hCoV-19/Norway/2813/2020\|EPI_ISL_493378\|2020-06-29 | ···················· | ·A·················· | ···················· | ···················· | ···················· | ···················· | |
| E | 8158 hCoV-19/Norway/2829/2020\|EPI_ISL_493382\|2020-06-29 | ···················· | ···················· | ···················· | ···················· | ···················· | ···················· | |
| E | 8159 hCoV-19/Norway/2831/2020\|EPI_ISL_493384\|2020-06-30 | ···················· | ···················· | ···················· | ···················· | ···················· | ···················· | |
| E | 8160 hCoV-19/Norway/2832/2020\|EPI_ISL_493385\|2020-06-30 | ···················· | ···················· | ···················· | ···················· | ···················· | ···················· | |
| E | 8161 hCoV-19/Norway/2834/2020\|EPI_ISL_493387\|2020-06-29 | ···················· | ·A·················· | ···················· | ···················· | ···················· | ···················· | |
| E | 8162 hCoV-19/Norway/2836/2020\|EPI_ISL_493389\|2020-06-30 | ···················· | ···················· | ···················· | ···················· | ···················· | ···················· | |
| E | 8163 hCoV-19/Singapore/630/2020\|EPI_ISL_493390\|2020-02- | ···················· | ···················· | ···················· | ···················· | ···················· | ···················· | |
| E | 8164 hCoV-19/Singapore/631/2020\|EPI_ISL_493391\|2020-04- | ···················· | ···················· | ···················· | ···················· | ···················· | ···················· | |
| E | 8165 hCoV-19/Singapore/632/2020\|EPI_ISL_493392\|2020-04- | ···················· | ···················· | ···················· | ···················· | ···················· | ···················· | |
| E | 8166 hCoV-19/Singapore/635/2020\|EPI_ISL_493395\|2020-04- | ···················· | ···················· | ···················· | ···················· | ··················NN | ···················· | 1 |
| E | 8167 hCoV-19/USA/CA-ALSR-1537/2020\|EPI_ISL_494650\|2020- | ···················· | ···················· | ···················· | ···················· | ···················· | ···················· | |
| E | 8168 hCoV-19/USA/CA-ALSR-1538/2020\|EPI_ISL_494651\|2020- | ···················· | ·A·················· | ···················· | ···················· | ···················· | ···················· | |
| E | 8169 hCoV-19/USA/CA-ALSR-1543/2020\|EPI_ISL_494652\|2020- | ···················· | ···················· | ···················· | ···················· | ···················· | ···················· | |
| E | 8170 hCoV-19/USA/CA-ALSR-1545/2020\|EPI_ISL_494654\|2020- | ···················· | ···················· | ···················· | ···················· | ···················· | ···················· | |
| E | 8171 hCoV-19/USA/CA-ALSR-1546/2020\|EPI_ISL_494655\|2020- | ···················· | ·A·················· | ···················· | ···················· | ···················· | ···················· | |
| E | 8172 hCoV-19/USA/CA-ALSR-1549/2020\|EPI_ISL_494656\|2020- | ···················· | ·A·················· | ···················· | ···················· | ···················· | ···················· | |
| E | 8173 hCoV-19/USA/CA-ALSR-1550/2020\|EPI_ISL_494657\|2020- | ···················· | ·A·················· | ···················· | ···················· | ···················· | ···················· | |
| E | 8174 hCoV-19/USA/CA-ALSR-1551/2020\|EPI_ISL_494658\|2020- | ···················· | ·A·················· | ···················· | ···················· | ···················· | ···················· | |
| E | 8175 hCoV-19/USA/CA-ALSR-1556/2020\|EPI_ISL_494659\|2020- | ···················· | ·A·················· | ···················· | ···················· | ···················· | ···················· | |
| E | 8176 hCoV-19/USA/CA-ALSR-1558/2020\|EPI_ISL_494660\|2020- | ···················· | ·A·················· | ···················· | ···················· | ···················· | ···················· | |
| E | 8177 hCoV-19/USA/CA-ALSR-1566/2020\|EPI_ISL_494662\|2020- | ···················· | ·A·················· | ···················· | ···················· | ···················· | ···················· | |
| E | 8178 hCoV-19/USA/CA-ALSR-1567/2020\|EPI_ISL_494663\|2020- | ···················· | ·A·················· | ···················· | ···················· | ···················· | ···················· | |
| E | 8179 hCoV-19/USA/CA-ALSR-1570/2020\|EPI_ISL_494664\|2020- | ···················· | ·A·················· | ···················· | ···················· | ···················· | ···················· | |
| E | 8180 hCoV-19/USA/CA-ALSR-1572/2020\|EPI_ISL_494665\|2020- | ···················· | ···················· | ···················· | ···················· | ···················· | ···················· | |
| E | 8181 hCoV-19/USA/CA-ALSR-1573/2020\|EPI_ISL_494666\|2020- | ···················· | ···················· | ···················· | ···················· | ···················· | ···················· | |
| E | 8182 hCoV-19/USA/CA-ALSR-1574/2020\|EPI_ISL_494667\|2020- | ···················· | ···················· | ···················· | ···················· | ···················· | ···················· | |
| E | 8183 hCoV-19/USA/CA-ALSR-1578/2020\|EPI_ISL_494668\|2020- | ···················· | ···················· | ···················· | ···················· | ···················· | ···················· | |
| E | 8184 hCoV-19/USA/CA-ALSR-1580/2020\|EPI_ISL_494669\|2020- | ···················· | ·A·················· | ···················· | ···················· | ···················· | ···················· | |
| E | 8185 hCoV-19/USA/CA-ALSR-1583/2020\|EPI_ISL_494671\|2020- | ···················· | ·A·················· | ···················· | ···················· | ···················· | ···················· | |
| E | 8186 hCoV-19/USA/CA-ALSR-1610/2020\|EPI_ISL_494685\|2020- | ···················· | ·A·················· | ···················· | ···················· | ···················· | ···················· | |
| E | 8187 hCoV-19/USA/CA-ALSR-1623/2020\|EPI_ISL_494694\|2020- | ···················· | ·A·················· | ···················· | ···················· | ···················· | ···················· | |
| E | 8188 hCoV-19/USA/CA-ALSR-1625/2020\|EPI_ISL_494695\|2020- | ···················· | ···················· | ···················· | ···················· | ···················· | ···················· | |
| E | 8189 hCoV-19/USA/CA-ALSR-1626/2020\|EPI_ISL_494696\|2020- | ···················· | ·A·················· | ···················· | ···················· | ···················· | ···················· | |
| E | 8190 hCoV-19/USA/CA-ALSR-1627/2020\|EPI_ISL_494697\|2020- | ···················· | ···················· | ···················· | ···················· | ···················· | ···················· | |
| E | 8191 hCoV-19/USA/CA-ALSR-1628/2020\|EPI_ISL_494698\|2020- | ···················· | ···················· | ···················· | ···················· | ···················· | ···················· | |
| E | 8192 hCoV-19/USA/CA-ALSR-1629/2020\|EPI_ISL_494699\|2020- | ···················· | ···················· | ···················· | ···················· | ···················· | ···················· | |
| E | 8193 hCoV-19/Japan/PG-0325/2020\|EPI_ISL_480000\|2020-03- | ···················· | ···················· | ···················· | ···················· | ···················· | ···················· | |
| E | 8194 hCoV-19/Japan/PG-0326/2020\|EPI_ISL_480001\|2020-03- | ···················· | ···················· | ···················· | ···················· | ···················· | ···················· | |
| E | 8195 hCoV-19/Japan/PG-0328/2020\|EPI_ISL_480003\|2020-03- | ···················· | ···················· | ···················· | ···················· | ···················· | ···················· | |
| E | 8196 hCoV-19/Japan/PG-0329/2020\|EPI_ISL_480004\|2020-03- | ···················· | ···················· | ···················· | ···················· | ···················· | ···················· | |
| E | 8197 hCoV-19/Japan/PG-0331/2020\|EPI_ISL_480005\|2020-03- | ···················· | ···················· | ···················· | ···················· | ···················· | ···················· | |
| E | 8198 hCoV-19/Japan/PG-0332/2020\|EPI_ISL_480006\|2020-03- | ···················· | ···················· | ···················· | ···················· | ···················· | ···················· | |
| E | 8199 hCoV-19/Japan/PG-0333/2020\|EPI_ISL_480007\|2020-03- | ···················· | ···················· | ···················· | ···················· | ···················· | ···················· | |
| E | 8200 hCoV-19/Japan/PG-0334/2020\|EPI_ISL_480008\|2020-03- | ···················· | ···················· | ···················· | ···················· | ···················· | ···················· | |
| E | 8201 hCoV-19/Japan/PG-0336/2020\|EPI_ISL_480009\|2020-03- | ···················· | ···················· | ···················· | ···················· | ···················· | ···················· | |
| E | 8202 hCoV-19/Japan/PG-0361/2020\|EPI_ISL_480031\|2020-02- | ···················· | ···················· | ···················· | ···················· | ···················· | ···················· | |
| E | 8203 hCoV-19/Japan/PG-0362/2020\|EPI_ISL_480032\|2020-02- | ···················· | ···················· | ···················· | ···················· | ···················· | ···················· | |
| E | 8204 hCoV-19/Japan/PG-0363/2020\|EPI_ISL_480033\|2020-02- | ···················· | ···················· | ···················· | ···················· | ···················· | ···················· | |
| E | 8205 hCoV-19/Japan/PG-0364/2020\|EPI_ISL_480034\|2020-03- | ···················· | ···················· | ···················· | ···················· | ···················· | ···················· | |
| E | 8206 hCoV-19/Japan/PG-0365/2020\|EPI_ISL_480035\|2020-03- | ···················· | ···················· | ···················· | ···················· | ···················· | ···················· | |
| E | 8207 hCoV-19/Japan/PG-0366/2020\|EPI_ISL_480036\|2020-03- | ···················· | ···················· | ···················· | ···················· | ···················· | ···················· | |
| E | 8208 hCoV-19/Japan/PG-0367/2020\|EPI_ISL_480037\|2020-03- | ···················· | ···················· | ···················· | ···················· | ···················· | ···················· | |

E 8209 hCoV-19/Japan/PG-0368/2020|EPI_ISL_480038|2020-03- ···················· ···················· ···················· ···················· ···················· ····················
E 8210 hCoV-19/Japan/PG-0369/2020|EPI_ISL_480039|2020-03- ···················· ···················· ···················· ···················· ···················· ····················
E 8211 hCoV-19/Japan/PG-0370/2020|EPI_ISL_480040|2020-03- ···················· ···················· ···················· ···················· ···················· ····················
E 8212 hCoV-19/Japan/PG-0371/2020|EPI_ISL_480041|2020-03- ···················· ···················· ···················· ···················· ···················· ····················
E 8213 hCoV-19/Japan/PG-0372/2020|EPI_ISL_480042|2020-02- ···················· ···················· ···················· ···················· ···················· ····················
E 8214 hCoV-19/Japan/PG-0373/2020|EPI_ISL_480043|2020-02- ···················· ···················· ···················· ···················· ···················· ····················
E 8215 hCoV-19/Japan/PG-0374/2020|EPI_ISL_480044|2020-02- ···················· ···················· ···················· ···················· ···················· ····················
E 8216 hCoV-19/Japan/PG-0375/2020|EPI_ISL_480045|2020-02- ···················· ···················· ···················· ···················· ···················· ····················
E 8217 hCoV-19/Japan/PG-0376/2020|EPI_ISL_480046|2020-02- ···················· ···················· ···················· ···················· ···················· ····················
E 8218 hCoV-19/Japan/PG-0377/2020|EPI_ISL_480047|2020-02- ···················· ···················· ···················· ···················· ···················· ····················
E 8219 hCoV-19/Japan/PG-0378/2020|EPI_ISL_480048|2020-02- ···················· ···················· ···················· ···················· ···················· ····················
E 8220 hCoV-19/Japan/PG-0379/2020|EPI_ISL_480049|2020-02- ···················· ···················· ···················· ···················· ···················· ····················
E 8221 hCoV-19/Japan/PG-0381/2020|EPI_ISL_480050|2020-03- ···················· ···················· ···················· ···················· ···················· ····················
E 8222 hCoV-19/Japan/PG-0382/2020|EPI_ISL_480051|2020-03- ···················· ···················· ···················· ···················· ···················· ····················
E 8223 hCoV-19/Japan/PG-0383/2020|EPI_ISL_480052|2020-03- ···················· ···················· ···················· ···················· ···················· ····················
E 8224 hCoV-19/South Korea/KCDC2050/2020|EPI_ISL_481370|2 ···················· ···················· ···················· ···················· ···················· ····················
E 8225 hCoV-19/South Korea/KCDC2051/2020|EPI_ISL_481371|2 ···················· ···················· ···················· ···················· ···················· ····················
E 8226 hCoV-19/South Korea/KCDC2052/2020|EPI_ISL_481372|2 ···················· ···················· ···················· ···················· ···················· ····················
E 8227 hCoV-19/South Korea/KCDC2053/2020|EPI_ISL_481373|2 ···················· ···················· ···················· ···················· ···················· ····················
E 8228 hCoV-19/South Korea/KCDC2054/2020|EPI_ISL_481374|2 ···················· ···················· ···················· ···················· ···················· ····················
E 8229 hCoV-19/South Korea/KCDC2055/2020|EPI_ISL_481375|2 ···················· ···················· ···················· ···················· ···················· ····················
E 8230 hCoV-19/South Korea/KCDC2056/2020|EPI_ISL_481376|2 ···················· ···················· ···················· ···················· ···················· ····················
E 8231 hCoV-19/South Korea/KCDC2057/2020|EPI_ISL_481377|2 ···················· ···················· ···················· ···················· ···················· ····················
E 8232 hCoV-19/South Korea/KCDC2058/2020|EPI_ISL_481378|2 ···················· ···················· ···················· ···················· ···················· ····················
E 8233 hCoV-19/South Korea/KCDC2059/2020|EPI_ISL_481379|2 ···················· ···················· ···················· ···················· ···················· ····················
E 8234 hCoV-19/Georgia/Tb-7856/2020|EPI_ISL_481380|2020-0 ···················· ···················· ···················· ···················· ···················· ····················
E 8235 hCoV-19/India/DL-NCDC5723-CSIR-IGIB/2020|EPI_ISL_4 ···················· ·R·················· ···················· ···················· ···················· ··N·················
E 8236 hCoV-19/India/DL-NCDC5725-CSIR-IGIB/2020|EPI_ISL_4 ···················· ·A·················· ···················· ···················· ···················· ··N·················
E 8237 hCoV-19/India/DL-NCDC5726-CSIR-IGIB/2020|EPI_ISL_4 ···················· ···················· ···················· ···················· ···················· ··N·················
E 8238 hCoV-19/India/DL-NCDC5743-CSIR-IGIB/2020|EPI_ISL_4 ···················· ·R·················· ···················· ···················· ···················· ··N·················
E 8239 hCoV-19/India/DL-NCDC6082-CSIR-IGIB/2020|EPI_ISL_4 ···················· ·A·················· ···················· ···················· ···················· ··A·················
E 8240 hCoV-19/India/DL-NCDC7032-CSIR-IGIB/2020|EPI_ISL_4 ···················· ···················· ···················· ···················· ···················· ····················
E 8241 hCoV-19/India/DL-NCDC7090-CSIR-IGIB/2020|EPI_ISL_4 ···················· ·A·················· ···················· ···················· ···················· ··A·················
E 8242 hCoV-19/India/DL-NCDC7101-CSIR-IGIB/2020|EPI_ISL_4 ···················· ·A·················· ···················· ···················· ···················N ··A················· 1
E 8243 hCoV-19/India/DL-NCDC7102-CSIR-IGIB/2020|EPI_ISL_4 ···················· ·A·················· ···················· ···················· ···················N ··A················· 1
E 8244 hCoV-19/India/DL-NCDC7104-CSIR-IGIB/2020|EPI_ISL_4 ···················· ·A·················· ···················· ···················· ········N··········· ··A·················
E 8245 hCoV-19/India/DL-NCDC7512-CSIR-IGIB/2020|EPI_ISL_4 ···················· ·N·················· ···················· ···················· ···················· ··R·················
E 8246 hCoV-19/India/DL-NCDC7515-CSIR-IGIB/2020|EPI_ISL_4 ···················· ·A·················· ···················· ···················· ···················· ··R·················
E 8247 hCoV-19/India/DL-NCDC7527-CSIR-IGIB/2020|EPI_ISL_4 ···················· ·N·················· ···················· ···················· ···················· ··R·················
E 8248 hCoV-19/India/DL-NCDC7549-CSIR-IGIB/2020|EPI_ISL_4 ···················· ·A·················· ···················· ···················· ···················· ··R·················
E 8249 hCoV-19/India/DL-NCDC7554-CSIR-IGIB/2020|EPI_ISL_4 ···················· ·A·················· ···················· ···················· ···················· ··A·················
E 8250 hCoV-19/India/DL-NCDC7556-CSIR-IGIB/2020|EPI_ISL_4 ···················· ·R·················· ···················· ···················· ···················· ····················
E 8251 hCoV-19/India/DL-NCDC7557-CSIR-IGIB/2020|EPI_ISL_4 ···················· ···················· ···················· ···················· ···················· ····················
E 8252 hCoV-19/India/DL-NCDC7624-CSIR-IGIB/2020|EPI_ISL_4 ···················· ···················· ···················· ···················· ···················· ····················
E 8253 hCoV-19/India/DL-NCDC7629-CSIR-IGIB/2020|EPI_ISL_4 ···················· ···················· ···················· ···················· ···················· ····················
E 8254 hCoV-19/India/DL-NCDC7649-CSIR-IGIB/2020|EPI_ISL_4 ···················· ···················· ···················· ···················· ···················· ··N·················
E 8255 hCoV-19/India/DL-NCDC7661-CSIR-IGIB/2020|EPI_ISL_4 ···················· ·R·················· ···················· ···················· ···················· ··R·················
E 8256 hCoV-19/India/DL-NCDC7686-CSIR-IGIB/2020|EPI_ISL_4 ···················· ···················· ···················· ···················· ···················· ····················
E 8257 hCoV-19/India/DL-NCDC7715-CSIR-IGIB/2020|EPI_ISL_4 ···················· ·R·················· ···················· ···················· ···················· ··R·················
E 8258 hCoV-19/India/DL-NCDC7717-CSIR-IGIB/2020|EPI_ISL_4 ···················· ···················· ···················· ···················· ···················· ··R·················
E 8259 hCoV-19/India/DL-NCDC7721-CSIR-IGIB/2020|EPI_ISL_4 ···················· ·R·················· ···················· ···················· ···················· ··N·················
E 8260 hCoV-19/India/DL-NCDC7728-CSIR-IGIB/2020|EPI_ISL_4 ···················· ···················· ···················· ···················· ···················· ····················
E 8261 hCoV-19/India/DL-NCDC7757-CSIR-IGIB/2020|EPI_ISL_4 ···················· ·A·················· ···················· ···················· ···················· ··R·················
E 8262 hCoV-19/Singapore/546/2020|EPI_ISL_482693|2020-05- ···················· ···················· ···················· ···················· ···················· ····················
E 8263 hCoV-19/Singapore/547/2020|EPI_ISL_482694|2020-05- ···················· ···················· ···················· ···················· ···················· ····················
E 8264 hCoV-19/Singapore/548/2020|EPI_ISL_482695|2020-05- ···················· ···················· ···················· ···················· ···················· ····················
E 8265 hCoV-19/Singapore/549/2020|EPI_ISL_482696|2020-05- ···················· ···················· ···················· ···················· ···················· ····················
E 8266 hCoV-19/Singapore/550/2020|EPI_ISL_482697|2020-05- ···················· ···················· ···················· ···················· ···················· ····················
E 8267 hCoV-19/Singapore/551/2020|EPI_ISL_482698|2020-05- ···················· ···················· ···················· ···················· ···················· ····················
E 8268 hCoV-19/Singapore/552/2020|EPI_ISL_482699|2020-06- ···················· ···················· ···················· ···················· ···················· ····················
E 8269 hCoV-19/England/GSTT-265D1D8/2020|EPI_ISL_483923|2 ···················· ···················· ···················· ···················· ···················· ····················
E 8270 hCoV-19/England/GSTT-265D25D/2020|EPI_ISL_483931|2 ···················· ···················· ···················· ···················· ···················· ····················
E 8271 hCoV-19/England/GSTT-265D2D5/2020|EPI_ISL_483937|2 ···················· ·A·················· ···················· ···················· ···················· ····················
E 8272 hCoV-19/England/GSTT-265D35A/2020|EPI_ISL_483945|2 ···················· ···················· ···················· ···················· ···················· ····················
E 8273 hCoV-19/England/GSTT-265D378/2020|EPI_ISL_483947|2 ···················· ···················· ···················· ···················· ···················· ····················
E 8274 hCoV-19/England/GSTT-265D387/2020|EPI_ISL_483948|2 ···················· ···················· ···················· ···················· ···················· ····················
E 8275 hCoV-19/England/GSTT-265D3C3/2020|EPI_ISL_483952|2 ···················· ···················· ···················· ···················· ···················· ····················
E 8276 hCoV-19/England/GSTT-265D439/2020|EPI_ISL_483959|2 ···················· ···················· ···················· ···················· ···················· ····················
E 8277 hCoV-19/England/GSTT-265D466/2020|EPI_ISL_483961|2 ···················· ···················· ···················· ···················· ···················· ····················
E 8278 hCoV-19/England/GSTT-265D493/2020|EPI_ISL_483964|2 ···················· ···················· ···················· ···················· ···················· ····················
E 8279 hCoV-19/England/GSTT-265D4B1/2020|EPI_ISL_483966|2 ···················· ···················· ···················· ···················· ···················· ····················
E 8280 hCoV-19/England/GSTT-265D4C0/2020|EPI_ISL_483967|2 ···················· ···················· ···················· ···················· ···················· ····················
E 8281 hCoV-19/England/GSTT-265D4DF/2020|EPI_ISL_483968|2 ···················· ···················· ···················· ···················· ···················· ····················
E 8282 hCoV-19/England/GSTT-265D509/2020|EPI_ISL_483971|2 ···················· ···················· ···················· ···················· ···················· ····················
E 8283 hCoV-19/England/GSTT-265D527/2020|EPI_ISL_483973|2 ···················· ···················· ···················· ···················· ···················· ····················
E 8284 hCoV-19/England/GSTT-265D554/2020|EPI_ISL_483976|2 ···················· ···················· ···················· ···················· ···················· ····················
E 8285 hCoV-19/England/GSTT-265D590/2020|EPI_ISL_483980|2 ···················· ···················· ···················· ···················· ···················· ····················
E 8286 hCoV-19/England/GSTT-265D5BE/2020|EPI_ISL_483982|2 ···················· ···················· ···················· ···················· ···················· ····················
E 8287 hCoV-19/England/GSTT-265D5EB/2020|EPI_ISL_483985|2 ···················· ···················· ···················· ···················· ···················· ····················
E 8288 hCoV-19/England/GSTT-265D5FA/2020|EPI_ISL_483986|2 ···················· ···················· ···················· ···················· ···················· ····················

| | | | | | | | | |
|---|---|---|---|---|---|---|---|---|
| E | 8289 hCoV-19/England/GSTT-265D606/2020\|EPI_ISL_483987\|2 | ···················· | ···················· | ···················· | ···················· | ···················· | ···················· | |
| E | 8290 hCoV-19/England/GSTT-265D624/2020\|EPI_ISL_483989\|2 | ···················· | ···················· | ···················· | ···················· | ···················· | ···················· | |
| E | 8291 hCoV-19/England/GSTT-265D651/2020\|EPI_ISL_483992\|2 | ···················· | ···················· | ···················· | ···················· | ···················· | ···················· | |
| E | 8292 hCoV-19/England/GSTT-265D68E/2020\|EPI_ISL_483995\|2 | ···················· | ···················· | ···················· | ···················· | ···················· | ···················· | |
| E | 8293 hCoV-19/England/GSTT-265D6CA/2020\|EPI_ISL_483999\|2 | ···················· | ···················· | ···················· | ···················· | ···················· | ···················· | |
| E | 8294 hCoV-19/USA/UT-02550/2020\|EPI_ISL_487203\|2020-04-2 | ···················· | ···················· | ···················· | ···················· | ···················· | ···················· | |
| E | 8295 hCoV-19/USA/UT-02556/2020\|EPI_ISL_487208\|2020-04-2 | ···················· | ···················· | ···················· | ···················· | ···················· | ···················· | |
| E | 8296 hCoV-19/USA/UT-02559/2020\|EPI_ISL_487210\|2020-04-2 | ···················· | ···················· | ···················· | ···················· | ···················· | ···················· | |
| E | 8297 hCoV-19/USA/UT-02563/2020\|EPI_ISL_487212\|2020-04-2 | ···················· | ···················· | ···················· | ···················· | ···················· | ···················· | |
| E | 8298 hCoV-19/USA/UT-04117/2020\|EPI_ISL_487248\|2020-05-1 | ···················· | ···················· | ···················· | ···················· | ···················· | ···················· | |
| E | 8299 hCoV-19/USA/UT-04127/2020\|EPI_ISL_487251\|2020-05-1 | ···················· | ···················· | ···················· | ···················· | ···················· | ···················· | |
| E | 8300 hCoV-19/USA/UT-04130/2020\|EPI_ISL_487252\|2020-05-1 | ···················· | ···················· | ···················· | ···················· | ···················· | ···················· | |
| E | 8301 hCoV-19/Peru/LIM-INS-002/2020\|EPI_ISL_487269\|2020- | ···················· | ···················· | ···················· | ···················· | ···················· | ···················· | |
| E | 8302 hCoV-19/Bahrain/340217593/2020\|EPI_ISL_487270\|2020 | ···················· | ···················· | ···················· | ···················· | ···················· | ···················· | |
| E | 8303 hCoV-19/Australia/VIC2119/2020\|EPI_ISL_487271\|2020 | ···················· | ·A·················· | ···················· | ···················· | ···················· | ··A················· | |
| E | 8304 hCoV-19/Bahrain/920267586/2020\|EPI_ISL_487272\|2020 | ···················· | ···················· | ···················· | ···················· | ···················· | ···················· | |
| E | 8305 hCoV-19/Bahrain/110003611/2020\|EPI_ISL_487273\|2020 | ···················· | ···················· | ···················· | ···················· | ···················· | ···················· | |
| E | 8306 hCoV-19/Bahrain/023232093/2020\|EPI_ISL_487274\|2020 | ···················· | ···················· | ···················· | ···················· | ···················· | ···················· | |
| E | 8307 hCoV-19/cat/Belgium/BE-MG-0320/2020\|EPI_ISL_487275 | ···················· | ···················· | ···················· | ···················· | ···················· | ···················· | |
| E | 8308 hCoV-19/South Africa/KRISP-0286/2020\|EPI_ISL_48727 | ···················· | ···················· | ···················· | ···················· | ···················· | ···················· | |
| E | 8309 hCoV-19/South Africa/KRISP-0290/2020\|EPI_ISL_48728 | ···················· | ···················· | ···················· | ···················· | ···················· | ···················· | |
| E | 8310 hCoV-19/South Africa/KRISP-0291/2020\|EPI_ISL_48728 | ···················· | ···················· | ···················· | ···················· | ···················· | ···················· | |
| E | 8311 hCoV-19/South Africa/KRISP-0300/2020\|EPI_ISL_48728 | ···················· | ···················· | ···················· | ···················· | ···················· | ···················· | |
| E | 8312 hCoV-19/England/NORT-295E31/2020\|EPI_ISL_488534\|20 | ···················· | ···················· | ···················· | ···················· | ···················· | ···················· | |
| E | 8313 hCoV-19/England/NORT-283522/2020\|EPI_ISL_488535\|20 | ···················· | ···················· | ···················· | ···················· | ···················· | ···················· | |
| E | 8314 hCoV-19/England/NORT-295639/2020\|EPI_ISL_488537\|20 | ···················· | ···················· | ···················· | ···················· | ···················· | ···················· | |
| E | 8315 hCoV-19/England/NORT-284E08/2020\|EPI_ISL_488538\|20 | ···················· | ···················· | ···················· | ···················· | ···················· | ···················· | |
| E | 8316 hCoV-19/England/NORT-283759/2020\|EPI_ISL_488539\|20 | ···················· | ···················· | ···················· | ···················· | ···················· | ···················· | |
| E | 8317 hCoV-19/England/NORT-29560C/2020\|EPI_ISL_488540\|20 | ···················· | ···················· | ···················· | ···················· | ···················· | ···················· | |
| E | 8318 hCoV-19/England/NORT-2842A2/2020\|EPI_ISL_488541\|20 | ···················· | ···················· | ···················· | ···················· | ···················· | ···················· | |
| E | 8319 hCoV-19/England/NORT-282417/2020\|EPI_ISL_488543\|20 | ···················· | ···················· | ···················· | ···················· | ···················· | ···················· | |
| E | 8320 hCoV-19/England/NORT-2827A5/2020\|EPI_ISL_488544\|20 | ···················· | ···················· | ···················· | ···················· | ···················· | ···················· | |
| E | 8321 hCoV-19/England/NORT-2831C4/2020\|EPI_ISL_488578\|20 | ···················· | ···················· | ···················· | ···················· | ···················· | ···················· | |
| E | 8322 hCoV-19/England/NORT-2821B6/2020\|EPI_ISL_488579\|20 | ···················· | ···················· | ···················· | ···················· | ···················· | ···················· | |
| E | 8323 hCoV-19/England/NORT-28448E/2020\|EPI_ISL_488580\|20 | ···················· | ····A··············· | ···················· | ···················· | ···················· | ···················· | |
| E | 8324 hCoV-19/England/NORT-2949B9/2020\|EPI_ISL_488581\|20 | ···················· | ···················· | ···················· | ···················· | ···················· | ···················· | |
| E | 8325 hCoV-19/England/NORT-294E8D/2020\|EPI_ISL_488582\|20 | ···················· | ···················· | ···················· | ···················· | ···················· | ···················· | |
| E | 8326 hCoV-19/England/NORT-283829/2020\|EPI_ISL_488583\|20 | ···················· | ···················· | ···················· | ···················· | ···················· | ···················· | |
| E | 8327 hCoV-19/England/NORT-295271/2020\|EPI_ISL_488584\|20 | ···················· | ···················· | ···················· | ···················· | ···················· | ···················· | |
| E | 8328 hCoV-19/England/NORT-294AC5/2020\|EPI_ISL_488585\|20 | ···················· | ···················· | ···················· | ···················· | ···················· | ···················· | |
| E | 8329 hCoV-19/England/NORT-283BA8/2020\|EPI_ISL_488586\|20 | ···················· | ···················· | ···················· | ···················· | ···················· | ···················· | |
| E | 8330 hCoV-19/England/NORT-283E54/2020\|EPI_ISL_488587\|20 | ···················· | ···················A | ···················· | ···················· | ···················· | ···················· | 1 |
| E | 8331 hCoV-19/England/NORT-2842EE/2020\|EPI_ISL_488588\|20 | ···················· | ···················· | ···················· | ···················· | ···················· | ···················· | |
| E | 8332 hCoV-19/England/NORT-283EDC/2020\|EPI_ISL_488589\|20 | ···················· | ···················· | ···················· | ···················· | ···················· | ···················· | |
| E | 8333 hCoV-19/England/NORT-283E18/2020\|EPI_ISL_488590\|20 | ···················· | ···················· | ···················· | ···················· | ···················· | ···················· | |
| E | 8334 hCoV-19/England/NORT-294D80/2020\|EPI_ISL_488591\|20 | ···················· | ···················· | ···················· | ···················· | ···················· | ···················· | |
| E | 8335 hCoV-19/England/NORT-295B49/2020\|EPI_ISL_488592\|20 | ···················· | ···················· | ···················· | ···················· | ···················· | ···················· | |
| E | 8336 hCoV-19/England/NORT-294F4E/2020\|EPI_ISL_488593\|20 | ···················· | ···················· | ···················· | ···················· | ···················· | ···················· | |
| E | 8337 hCoV-19/England/NORT-2829EB/2020\|EPI_ISL_488594\|20 | ···················· | ···················· | ···················· | ···················· | ···················· | ···················· | |
| E | 8338 hCoV-19/England/NORT-295B58/2020\|EPI_ISL_488595\|20 | ···················· | ···················· | ···················· | ···················· | ···················· | ···················· | |
| E | 8339 hCoV-19/England/NORT-294C29/2020\|EPI_ISL_488596\|20 | ···················· | ···················· | ···················· | ···················· | ···················· | ···················· | |
| E | 8340 hCoV-19/England/NORT-295A5B/2020\|EPI_ISL_488598\|20 | ···················· | ···················· | ···················· | ···················· | ···················· | ···················· | |
| E | 8341 hCoV-19/England/NORT-294A89/2020\|EPI_ISL_488599\|20 | ···················· | ···················· | ···················· | ···················· | ···················· | ···················· | |
| E | 8342 hCoV-19/Wales/PHWC-1662E3/2020\|EPI_ISL_490861\|2020 | ···················· | ···················· | ···················· | ···················· | ···················· | ···················· | |
| E | 8343 hCoV-19/Wales/PHWC-1662F2/2020\|EPI_ISL_490862\|2020 | ···················· | ···················· | ···················· | ···················· | ···················· | ···················· | |
| E | 8344 hCoV-19/Wales/PHWC-16632C/2020\|EPI_ISL_490865\|2020 | ···················· | ···················· | ···················· | ···················· | ···················· | ···················· | |
| E | 8345 hCoV-19/Wales/PHWC-16633B/2020\|EPI_ISL_490866\|2020 | ···················· | ···················· | ···················· | ···················· | ···················· | ···················· | |
| E | 8346 hCoV-19/Wales/PHWC-16634A/2020\|EPI_ISL_490867\|2020 | ···················· | ···················· | ···················· | ···················· | ···················· | ···················· | |
| E | 8347 hCoV-19/Oman/205040055/2020\|EPI_ISL_492010\|2020-05 | ···················· | ···················· | ···················· | ···················· | ···················· | ···················· | |
| E | 8348 hCoV-19/Oman/205040106/2020\|EPI_ISL_492011\|2020-05 | ···················· | ···················· | ···················· | ···················· | ···················· | ···················· | |
| E | 8349 hCoV-19/Oman/205040194/2020\|EPI_ISL_492012\|2020-05 | ···················· | ···················· | ···················· | ···················· | ···················· | ···················· | |
| E | 8350 hCoV-19/Oman/205041214/2020\|EPI_ISL_492014\|2020-06 | ···················· | ···················· | ···················· | ···················· | ···················· | ···················· | |
| E | 8351 hCoV-19/Oman/205041374/2020\|EPI_ISL_492016\|2020-06 | ···················· | ···················· | ···················· | ···················· | ···················· | ···················· | |
| E | 8352 hCoV-19/Oman/205041592/2020\|EPI_ISL_492017\|2020-06 | ···················· | ·A·················· | ···················· | ···················· | ···················· | ···················· | |
| E | 8353 hCoV-19/Oman/205042592/2020\|EPI_ISL_492019\|2020-06 | ···················· | ···················· | ···················· | ···················· | ···················· | ···················· | |
| E | 8354 hCoV-19/Oman/205042822/2020\|EPI_ISL_492020\|2020-06 | ···················· | ···················· | ···················· | ···················· | ···················· | ···················· | |
| E | 8355 hCoV-19/Oman/205042904/2020\|EPI_ISL_492021\|2020-06 | ···················· | ···················· | ···················· | ···················· | ···················· | ···················· | |
| E | 8356 hCoV-19/Oman/205042917/2020\|EPI_ISL_492022\|2020-06 | ···················· | ···················· | ···················· | ···················· | ···················· | ···················· | |
| E | 8357 hCoV-19/Oman/205042975/2020\|EPI_ISL_492023\|2020-06 | ···················· | ·A·················· | ···················· | ···················· | ···················· | ···················· | |
| E | 8358 hCoV-19/Oman/205043002/2020\|EPI_ISL_492024\|2020-06 | ···················· | ···················· | ···················· | ···················· | ···················· | ···················· | |
| E | 8359 hCoV-19/Oman/205043071/2020\|EPI_ISL_492025\|2020-06 | ···················· | ···················· | ···················· | ···················· | ···················· | ···················· | |
| E | 8360 hCoV-19/Oman/205043490/2020\|EPI_ISL_492026\|2020-06 | ···················· | ·A·················· | ···················· | ···················· | ···················· | ···················· | |
| E | 8361 hCoV-19/Bangladesh/CHRF-0018/2020\|EPI_ISL_492029\|2 | ···················· | ···················· | ···················· | ···················· | ···················· | ···················· | |
| E | 8362 hCoV-19/Bangladesh/CHRF-0019/2020\|EPI_ISL_492030\|2 | ···················· | ···················· | ···················· | ···················· | ···················· | ···················· | |
| E | 8364 hCoV-19/Mexico/YUC-InDRE_33/2020\|EPI_ISL_493348\|20 | ···················· | ···················· | ···················· | ···················· | ···················· | ···················· | |
| E | 8365 hCoV-19/Norway/2781/2020\|EPI_ISL_493352\|2020-06-22 | ···················· | ···················· | ···················· | ···················· | ···················· | ···················· | |
| E | 8366 hCoV-19/Norway/2782/2020\|EPI_ISL_493353\|2020-06-22 | ···················· | ···················· | ···················· | ···················· | ···················· | ···················· | |
| E | 8367 hCoV-19/Norway/2784/2020\|EPI_ISL_493354\|2020-06-23 | ···················· | ···················· | ···················· | ···················· | ···················· | ···················· | |
| E | 8368 hCoV-19/Norway/2785/2020\|EPI_ISL_493355\|2020-06-23 | ···················· | ···················· | ···················· | ···················· | ···················· | ···················· | |
| E | 8369 hCoV-19/Norway/2789/2020\|EPI_ISL_493356\|2020-06-26 | ···················· | ·A·················· | ···················· | ···················· | ···················· | ···················· | |

| | | | | | | | |
|---|---|---|---|---|---|---|---|
| E | 8370 hCoV-19/Norway/2790/2020\|EPI_ISL_493357\|2020-06-26 | ···················· | ·A·················· | ····················· | ···················· | ···················· | ···················· |
| E | 8371 hCoV-19/Norway/2792/2020\|EPI_ISL_493358\|2020-06-23 | ···················· | ···················· | ····················· | ···················· | ···················· | ···················· |
| E | 8372 hCoV-19/Norway/2793/2020\|EPI_ISL_493359\|2020-06-25 | ···················· | ·A·················· | ····················· | ···················· | ···················· | ···················· |
| E | 8373 hCoV-19/Norway/2794/2020\|EPI_ISL_493360\|2020-06-25 | ···················· | ·A·················· | ····················· | ···················· | ···················· | ···················· |
| E | 8374 hCoV-19/Norway/2796/2020\|EPI_ISL_493362\|2020-06-25 | ···················· | ···················· | ····················· | ···················· | ···················· | ···················· |
| E | 8375 hCoV-19/USA/CA-ALSR-1435/2020\|EPI_ISL_494601\|2020- | ···················· | ···················· | ····················· | ···················· | ···················· | ···················· |
| E | 8376 hCoV-19/USA/CA-ALSR-1437/2020\|EPI_ISL_494603\|2020- | ···················· | ···················· | ····················· | ···················· | ···················· | ···················· |
| E | 8377 hCoV-19/USA/CA-ALSR-1441/2020\|EPI_ISL_494604\|2020- | ···················· | ···················· | ····················· | ···················· | ···················· | ···················· |
| E | 8378 hCoV-19/USA/CA-ALSR-1443/2020\|EPI_ISL_494605\|2020- | ···················· | ···················· | ····················· | ···················· | ···················· | ···················· |
| E | 8379 hCoV-19/USA/CA-ALSR-1446/2020\|EPI_ISL_494607\|2020- | ···················· | ···················· | ····················· | ···················· | ···················· | ···················· |
| E | 8380 hCoV-19/USA/CA-ALSR-1447/2020\|EPI_ISL_494608\|2020- | ···················· | ···················· | ····················· | ···················· | ···················· | ···················· |
| E | 8381 hCoV-19/USA/CA-ALSR-1448/2020\|EPI_ISL_494609\|2020- | ···················· | ···················· | ····················· | ···················· | ···················· | ···················· |
| E | 8382 hCoV-19/USA/CA-ALSR-1449/2020\|EPI_ISL_494610\|2020- | ···················· | ···················· | ····················· | ···················· | ···················· | ···················· |
| E | 8383 hCoV-19/USA/CA-ALSR-1451/2020\|EPI_ISL_494611\|2020- | ···················· | ···················· | ····················· | ···················· | ···················· | ···················· |
| E | 8384 hCoV-19/USA/CA-ALSR-1452/2020\|EPI_ISL_494612\|2020- | ···················· | ···················· | ····················· | ···················· | ···················· | ···················· |
| E | 8385 hCoV-19/USA/CA-ALSR-1453/2020\|EPI_ISL_494613\|2020- | ···················· | ···················· | ····················· | ···················· | ···················· | ···················· |
| E | 8386 hCoV-19/USA/CA-ALSR-1456/2020\|EPI_ISL_494615\|2020- | ···················· | ···················· | ····················· | ···················· | ···················· | ···················· |
| E | 8387 hCoV-19/USA/CA-ALSR-1461/2020\|EPI_ISL_494617\|2020- | ···················· | ···················· | ····················· | ···················· | ···················· | ···················· |
| E | 8388 hCoV-19/USA/CA-ALSR-1462/2020\|EPI_ISL_494618\|2020- | ···················· | ·············A······ | ····················· | ···················· | ···················· | ···················· |
| E | 8389 hCoV-19/USA/CA-ALSR-1463/2020\|EPI_ISL_494619\|2020- | ···················· | ···················· | ····················· | ···················· | ···················· | ···················· |
| E | 8390 hCoV-19/USA/CA-ALSR-1491/2020\|EPI_ISL_494620\|2020- | ···················· | ···················· | ····················· | ···················· | ···················· | ···················· |
| E | 8391 hCoV-19/USA/CA-ALSR-1495/2020\|EPI_ISL_494621\|2020- | ···················· | ·A·················· | ····················· | ···················· | ···················· | ···················· |
| E | 8392 hCoV-19/USA/CA-ALSR-1497/2020\|EPI_ISL_494622\|2020- | ···················· | ···················· | ····················· | ···················· | ···················· | ···················· |
| E | 8393 hCoV-19/USA/CA-ALSR-1498/2020\|EPI_ISL_494623\|2020- | ···················· | ···················· | ····················· | ···················· | ···················· | ···················· |
| E | 8394 hCoV-19/USA/CA-ALSR-1499/2020\|EPI_ISL_494624\|2020- | ···················· | ···················· | ····················· | ···················· | ···················· | ···················· |
| E | 8395 hCoV-19/USA/CA-ALSR-1502/2020\|EPI_ISL_494625\|2020- | ···················· | ·A·················· | ····················· | ···················· | ···················· | ···················· |
| E | 8396 hCoV-19/USA/CA-ALSR-1503/2020\|EPI_ISL_494626\|2020- | ···················· | ···················· | ····················· | ···················· | ···················· | ···················· |
| E | 8397 hCoV-19/USA/CA-ALSR-1504/2020\|EPI_ISL_494627\|2020- | ···················· | ···················· | ····················· | ···················· | ···················· | ···················· |
| E | 8398 hCoV-19/USA/CA-ALSR-1505/2020\|EPI_ISL_494628\|2020- | ···················· | ···················· | ····················· | ···················· | ···················· | ···················· |
| E | 8399 hCoV-19/USA/CA-ALSR-1506/2020\|EPI_ISL_494629\|2020- | ···················· | ···················· | ····················· | ···················· | ···················· | ···················· |
| E | 8400 hCoV-19/USA/CA-ALSR-1508/2020\|EPI_ISL_494630\|2020- | ···················· | ···················· | ····················· | ···················· | ···················· | ···················· |
| E | 8401 hCoV-19/USA/CA-ALSR-1510/2020\|EPI_ISL_494632\|2020- | ···················· | ···················· | ····················· | ···················· | ···················· | ···················· |
| E | 8402 hCoV-19/USA/CA-ALSR-1512/2020\|EPI_ISL_494634\|2020- | ···················· | ·A·················· | ····················· | ···················· | ···················· | ···················· |
| E | 8403 hCoV-19/USA/CA-ALSR-1513/2020\|EPI_ISL_494635\|2020- | ···················· | ·A·················· | ····················· | ···················· | ···················· | ···················· |
| E | 8404 hCoV-19/USA/CA-ALSR-1514/2020\|EPI_ISL_494636\|2020- | ···················· | ·A·················· | ····················· | ···················· | ···················· | ···················· |
| E | 8405 hCoV-19/USA/CA-ALSR-1517/2020\|EPI_ISL_494639\|2020- | ···················· | ·A·················· | ····················· | ···················· | ···················· | ···················· |
| E | 8406 hCoV-19/USA/CA-ALSR-1522/2020\|EPI_ISL_494641\|2020- | ···················· | ·A·················· | ····················· | ···················· | ···················· | ···················· |
| E | 8407 hCoV-19/USA/CA-ALSR-1525/2020\|EPI_ISL_494642\|2020- | ···················· | ·A·················· | ····················· | ···················· | ···················· | ···················· |
| E | 8408 hCoV-19/USA/CA-ALSR-1527/2020\|EPI_ISL_494644\|2020- | ···················· | ···················· | ····················· | ···················· | ···················· | ···················· |
| E | 8409 hCoV-19/USA/CA-ALSR-1530/2020\|EPI_ISL_494645\|2020- | ···················· | ···················· | ····················· | ···················· | ···················· | ···················· |
| E | 8410 hCoV-19/USA/CA-ALSR-1531/2020\|EPI_ISL_494646\|2020- | ···················· | ···················· | ····················· | ···················· | ···················· | ···················· |
| E | 8411 hCoV-19/USA/CA-ALSR-1533/2020\|EPI_ISL_494647\|2020- | ···················· | ·A·················· | ····················· | ···················· | ···················· | ···················· |
| E | 8412 hCoV-19/USA/CA-ALSR-1536/2020\|EPI_ISL_494649\|2020- | ···················· | ·A·················· | ····················· | ···················· | ···················· | ···················· |
| E | 8413 hCoV-19/USA/CA-ALSR-1584/2020\|EPI_ISL_494672\|2020- | ···················· | ·A·················· | ····················· | ···················· | ···················· | ···················· |
| E | 8414 hCoV-19/USA/CA-ALSR-1585/2020\|EPI_ISL_494673\|2020- | ···················· | ·A·················· | ····················· | ···················· | ···················· | ···················· |
| E | 8415 hCoV-19/USA/CA-ALSR-1586/2020\|EPI_ISL_494674\|2020- | ···················· | ···················· | ····················· | ···················· | ···················· | ···················· |
| E | 8416 hCoV-19/USA/CA-ALSR-1587/2020\|EPI_ISL_494675\|2020- | ···················· | ···················· | ····················· | ···················· | ···················· | ···················· |
| E | 8417 hCoV-19/USA/CA-ALSR-1588/2020\|EPI_ISL_494676\|2020- | ···················· | ···················· | ····················· | ···················· | ···················· | ···················· |
| E | 8418 hCoV-19/USA/CA-ALSR-1590/2020\|EPI_ISL_494678\|2020- | ···················· | ···················· | ····················· | ···················· | ···················· | ···················· |
| E | 8419 hCoV-19/USA/CA-ALSR-1593/2020\|EPI_ISL_494680\|2020- | ···················· | ···················· | ····················· | ···················· | ···················· | ···················· |
| E | 8420 hCoV-19/USA/CA-ALSR-1595/2020\|EPI_ISL_494681\|2020- | ···················· | ···················· | ····················· | ···················· | ···················· | ···················· |
| E | 8421 hCoV-19/USA/CA-ALSR-1596/2020\|EPI_ISL_494682\|2020- | ···················· | ·A·················· | ····················· | ···················· | ···················· | ···················· |
| E | 8422 hCoV-19/USA/CA-ALSR-1597/2020\|EPI_ISL_494683\|2020- | ···················· | ·A·················· | ····················· | ···················· | ···················· | ···················· |
| E | 8423 hCoV-19/USA/CA-ALSR-1603/2020\|EPI_ISL_494684\|2020- | ···················· | ···················· | ····················· | ···················· | ···················· | ···················· |
| E | 8424 hCoV-19/USA/CA-ALSR-1612/2020\|EPI_ISL_494686\|2020- | ···················· | ·A·················· | ····················· | ···················· | ···················· | ···················· |
| E | 8425 hCoV-19/USA/CA-ALSR-1614/2020\|EPI_ISL_494687\|2020- | ···················· | ·A·················· | ····················· | ···················· | ···················· | ···················· |
| E | 8426 hCoV-19/USA/CA-ALSR-1615/2020\|EPI_ISL_494688\|2020- | ···················· | ···················· | ····················· | ···················· | ···················· | ···················· |
| E | 8427 hCoV-19/USA/CA-ALSR-1616/2020\|EPI_ISL_494689\|2020- | ···················· | ···················· | ····················· | ···················· | ···················· | ···················· |
| E | 8428 hCoV-19/USA/CA-ALSR-1619/2020\|EPI_ISL_494690\|2020- | ···················· | ···················· | ····················· | ···················· | ···················· | ···················· |
| E | 8429 hCoV-19/USA/CA-ALSR-1620/2020\|EPI_ISL_494691\|2020- | ···················· | ···················· | ····················· | ···················· | ···················· | ···················· |
| E | 8430 hCoV-19/USA/CA-ALSR-1621/2020\|EPI_ISL_494692\|2020- | ···················· | ···················· | ····················· | ···················· | ···················· | ···················· |
| E | 8431 hCoV-19/USA/CA-ALSR-1622/2020\|EPI_ISL_494693\|2020- | ···················· | ·A·················· | ····················· | ···················· | ···················· | ···················· |
| E | 8432 hCoV-19/USA/WA-S1451/2020\|EPI_ISL_495901\|2020-05-2 | ···················· | ···················· | ····················· | ···················· | ···················· | ···················· |
| E | 8433 hCoV-19/USA/WA-S1452/2020\|EPI_ISL_495902\|2020-05-2 | ···················· | ···················· | ····················· | ···················· | ···················· | ···················· |
| E | 8434 hCoV-19/USA/WA-S1453/2020\|EPI_ISL_495903\|2020-05-2 | ···················· | ···················· | ····················· | ···················· | ···················· | ···················· |
| E | 8435 hCoV-19/USA/WA-S1454/2020\|EPI_ISL_495904\|2020-05-2 | ···················· | ···················· | ····················· | ···················· | ···················· | ···················· |
| E | 8436 hCoV-19/USA/WA-S1455/2020\|EPI_ISL_495905\|2020-05-2 | ···················· | ···················· | ····················· | ···················· | ···················· | ···················· |
| E | 8437 hCoV-19/USA/WA-S1456/2020\|EPI_ISL_495906\|2020-05-2 | ···················· | ···················· | ····················· | ···················· | ···················· | ···················· |
| E | 8438 hCoV-19/USA/WA-S1457/2020\|EPI_ISL_495907\|2020-05-2 | ···················· | ···················· | ····················· | ···················· | ···················· | ···················· |
| E | 8439 hCoV-19/USA/WA-S1458/2020\|EPI_ISL_495908\|2020-05-2 | ···················· | ···················· | ····················· | ···················· | ···················· | ···················· |
| E | 8440 hCoV-19/USA/WA-S1459/2020\|EPI_ISL_495909\|2020-05-2 | ···················· | ···················· | ····················· | ···················· | ···················· | ···················· |
| E | 8441 hCoV-19/USA/WA-S1460/2020\|EPI_ISL_495910\|2020-05-2 | ···················· | ···················· | ····················· | ···················· | ···················· | ···················· |
| E | 8442 hCoV-19/USA/WA-S1461/2020\|EPI_ISL_495911\|2020-05-2 | ···················· | ···················· | ····················· | ···················· | ···················· | ···················· |
| E | 8443 hCoV-19/USA/WA-S1462/2020\|EPI_ISL_495912\|2020-05-2 | ···················· | ···················· | ····················· | ···················· | ···················· | ···················· |
| E | 8444 hCoV-19/USA/WA-S1463/2020\|EPI_ISL_495913\|2020-05-2 | ···················· | ···················· | ····················· | ···················· | ···················· | ···················· |
| E | 8445 hCoV-19/USA/WA-S1464/2020\|EPI_ISL_495914\|2020-05-2 | ···················· | ···················· | ····················· | ···················· | ···················· | ···················· |
| E | 8446 hCoV-19/USA/WA-S1465/2020\|EPI_ISL_495915\|2020-05-2 | ···················· | ···················· | ····················· | ···················· | ···················· | ···················· |
| E | 8447 hCoV-19/USA/WA-S1466/2020\|EPI_ISL_495916\|2020-05-2 | ···················· | ···················· | ····················· | ···················· | ···················· | ···················· |
| E | 8448 hCoV-19/Bangladesh/BCSIR-NILMRC-254/2020\|EPI_ISL_5 | ···················· | ···················· | ····················· | ···················· | ···················· | ···················· |
| E | 8483 hCoV-19/Spain/MD-IBV-004943/2020\|EPI_ISL_481092\|20 | ···················· | ···················· | ····················· | ···················· | ···················· | ···················· |

| | | | | | | | | |
|---|---|---|---|---|---|---|---|---|
| E | 8490 | hCoV-19/USA/OR-PROV-079/2020\|EPI_ISL_482352\|2020-0 | .................... | .................... | .................... | .................... | .................... | .................... |
| E | 8491 | hCoV-19/USA/OR-PROV-080/2020\|EPI_ISL_482353\|2020-0 | .................... | .................... | .................... | .................... | .................... | .................... |
| E | 8492 | hCoV-19/USA/OR-PROV-081/2020\|EPI_ISL_482354\|2020-0 | .................... | .................... | .................... | .................... | .................... | .................... |
| E | 8493 | hCoV-19/USA/OR-PROV-083/2020\|EPI_ISL_482355\|2020-0 | .................... | .................... | .................... | .................... | .................... | .................... |
| E | 8494 | hCoV-19/USA/OR-PROV-085/2020\|EPI_ISL_482356\|2020-0 | .................... | .................... | .................... | .................... | .................... | .................... |
| E | 8495 | hCoV-19/USA/OR-PROV-086/2020\|EPI_ISL_482357\|2020-0 | .................... | .................... | .................... | .................... | .................... | .................... |
| E | 8496 | hCoV-19/USA/OR-PROV-087/2020\|EPI_ISL_482358\|2020-0 | .................... | .................... | .................... | .................... | .................... | .................... |
| E | 8497 | hCoV-19/USA/OR-PROV-088/2020\|EPI_ISL_482359\|2020-0 | .................... | .................... | .................... | .................... | .................... | .................... |
| E | 8498 | hCoV-19/USA/OR-PROV-089/2020\|EPI_ISL_482360\|2020-0 | .................... | .................... | .................... | .................... | .................... | .................... |
| E | 8499 | hCoV-19/USA/OR-PROV-090/2020\|EPI_ISL_482361\|2020-0 | .................... | .................... | .................... | .................... | .................... | .................... |
| E | 8500 | hCoV-19/USA/OR-PROV-091/2020\|EPI_ISL_482362\|2020-0 | .................... | .................... | .................... | .................... | .................... | .................... |
| E | 8501 | hCoV-19/USA/OR-PROV-133/2020\|EPI_ISL_482396\|2020-0 | .................... | .................... | .................... | .................... | .................... | .................... |
| E | 8502 | hCoV-19/USA/OR-PROV-135/2020\|EPI_ISL_482397\|2020-0 | .................... | .................... | .................... | .................... | .................... | .................... |
| E | 8503 | hCoV-19/USA/OR-PROV-136/2020\|EPI_ISL_482398\|2020-0 | .................... | .................... | .................... | .................... | .................... | .................... |
| E | 8504 | hCoV-19/USA/OR-PROV-137/2020\|EPI_ISL_482399\|2020-0 | .................... | .................... | .................... | .................... | .................... | .................... |
| E | 8506 | hCoV-19/Bangladesh/BCSIR-NILMRC-204/2020\|EPI_ISL_4 | .................... | .A.................. | .................... | .................... | .................... | ..A................. |
| E | 8507 | hCoV-19/Bangladesh/BCSIR-NILMRC-205/2020\|EPI_ISL_4 | .................... | .................... | .................... | .................... | .................... | .................... |
| E | 8508 | hCoV-19/Bangladesh/BCSIR-NILMRC-206/2020\|EPI_ISL_4 | .................... | .................... | .................... | .................... | .................... | .................... |
| E | 8509 | hCoV-19/Bangladesh/BCSIR-NILMRC-207/2020\|EPI_ISL_4 | .................... | .................... | .................... | .................... | .................... | .................... |
| E | 8510 | hCoV-19/Bangladesh/BCSIR-NILMRC-208/2020\|EPI_ISL_4 | .................... | .................... | .................... | .................... | .................... | .................... |
| E | 8511 | hCoV-19/Bangladesh/BCSIR-NILMRC-210/2020\|EPI_ISL_4 | .................... | .................... | .................... | .................... | .................... | .................... |
| E | 8512 | hCoV-19/Bangladesh/BCSIR-NILMRC-217/2020\|EPI_ISL_4 | .................... | .................... | .................... | .................... | .................... | .................... |
| E | 8513 | hCoV-19/Bangladesh/BCSIR-NILMRC-180/2020\|EPI_ISL_4 | .................... | .................... | .................... | .................... | .................... | .................... |
| E | 8514 | hCoV-19/USA/UT-02543/2020\|EPI_ISL_487200\|2020-04-2 | .................... | .................... | .................... | .................... | .................... | .................... |
| E | 8515 | hCoV-19/USA/UT-02554/2020\|EPI_ISL_487206\|2020-04-2 | .................... | .................... | .................... | .................... | .................... | .................... |
| E | 8516 | hCoV-19/USA/UT-02566/2020\|EPI_ISL_487215\|2020-04-2 | .................... | .................... | .................... | .................... | .................... | .................... |
| E | 8517 | hCoV-19/USA/UT-02571/2020\|EPI_ISL_487218\|2020-04-2 | .................... | .................... | .................... | .................... | .................... | .................... |
| E | 8518 | hCoV-19/USA/UT-02573/2020\|EPI_ISL_487219\|2020-04-2 | .................... | .................... | .................... | .................... | .................... | .................... |
| E | 8519 | hCoV-19/USA/UT-02574/2020\|EPI_ISL_487220\|2020-04-2 | .................... | .................... | .................... | .................... | .................... | .................... |
| E | 8520 | hCoV-19/USA/UT-02577/2020\|EPI_ISL_487221\|2020-04-2 | .................... | .................... | .................... | .................... | .................... | .................... |
| E | 8521 | hCoV-19/USA/UT-02581/2020\|EPI_ISL_487223\|2020-04-2 | .................... | .................... | .................... | .................... | .................... | .................... |
| E | 8522 | hCoV-19/USA/UT-02585/2020\|EPI_ISL_487225\|2020-04-2 | .................... | .................... | .................... | .................... | .................... | .................... |
| E | 8523 | hCoV-19/USA/UT-02588/2020\|EPI_ISL_487226\|2020-04-2 | .................... | .................... | .................... | .................... | .................... | .................... |
| E | 8525 | hCoV-19/USA/UT-04115/2020\|EPI_ISL_487246\|2020-05-1 | .................... | .................... | .................... | .................... | .................... | .................... |
| E | 8526 | hCoV-19/England/NORT-2955D2/2020\|EPI_ISL_488501\|20 | .................... | .................... | .................... | .................... | .................... | .................... |
| E | 8527 | hCoV-19/England/NORT-283B4E/2020\|EPI_ISL_488502\|20 | .................... | .................... | .................... | .................... | .................... | .................... |
| E | 8528 | hCoV-19/England/NORT-282E37/2020\|EPI_ISL_488503\|20 | .................... | .................... | .................... | .................... | .................... | .................... |
| E | 8529 | hCoV-19/England/NORT-283B3F/2020\|EPI_ISL_488504\|20 | .................... | .................... | .................... | .................... | .................... | .................... |
| E | 8530 | hCoV-19/England/NORT-283C87/2020\|EPI_ISL_488505\|20 | .................... | .................... | .................... | .................... | .................... | .................... |
| E | 8531 | hCoV-19/England/NORT-28267B/2020\|EPI_ISL_488506\|20 | .................... | .................... | .................... | .................... | .................... | .................... |
| E | 8532 | hCoV-19/England/NORT-294A10/2020\|EPI_ISL_488507\|20 | .................... | .................... | .................... | .................... | .................... | .................... |
| E | 8533 | hCoV-19/England/NORT-284293/2020\|EPI_ISL_488508\|20 | .................... | .................... | .................... | .................... | .................... | .................... |
| E | 8534 | hCoV-19/England/NORT-295DAD/2020\|EPI_ISL_488509\|20 | .................... | .................... | .................... | .................... | .................... | .................... |
| E | 8535 | hCoV-19/England/NORT-284794/2020\|EPI_ISL_488510\|20 | .................... | .................... | .................... | .................... | .................... | .................... |
| E | 8536 | hCoV-19/England/NORT-284CB3/2020\|EPI_ISL_488511\|20 | .................... | .................... | .................... | .................... | .................... | .................... |
| E | 8537 | hCoV-19/England/NORT-284CFF/2020\|EPI_ISL_488512\|20 | .................... | .................... | .................... | .................... | .................... | .................... |
| E | 8538 | hCoV-19/England/NORT-282DFE/2020\|EPI_ISL_488513\|20 | .................... | .................... | .................... | .................... | .................... | .................... |
| E | 8539 | hCoV-19/England/NORT-282ABB/2020\|EPI_ISL_488514\|20 | .................... | .................... | .................... | .................... | .................... | .................... |
| E | 8540 | hCoV-19/England/NORT-2953D8/2020\|EPI_ISL_488515\|20 | .................... | .................... | .................... | .................... | .................... | .................... |
| E | 8541 | hCoV-19/England/NORT-295851/2020\|EPI_ISL_488516\|20 | .................... | .................... | .................... | .................... | .................... | .................... |
| E | 8542 | hCoV-19/England/NORT-282E19/2020\|EPI_ISL_488517\|20 | .................... | .................... | .................... | .................... | .................... | .................... |
| E | 8543 | hCoV-19/England/NORT-282329/2020\|EPI_ISL_488518\|20 | .................... | .................... | .................... | .................... | .................... | .................... |
| E | 8544 | hCoV-19/England/NORT-2844AC/2020\|EPI_ISL_488519\|20 | .................... | .................... | .................... | .................... | .................... | .................... |
| E | 8545 | hCoV-19/England/NORT-2843BE/2020\|EPI_ISL_488520\|20 | .................... | .................... | .................... | .................... | .................... | .................... |
| E | 8546 | hCoV-19/England/NORT-295E04/2020\|EPI_ISL_488521\|20 | .................... | .................... | .................... | .................... | .................... | .................... |
| E | 8547 | hCoV-19/England/NORT-283A7E/2020\|EPI_ISL_488522\|20 | .................... | .................... | .................... | .................... | .................... | .................... |
| E | 8548 | hCoV-19/England/NORT-29550F/2020\|EPI_ISL_488523\|20 | .................... | .................... | .................... | .................... | .................... | .................... |
| E | 8549 | hCoV-19/England/NORT-2956EE/2020\|EPI_ISL_488524\|20 | .................... | .................... | .................... | .................... | .................... | .................... |
| E | 8550 | hCoV-19/England/NORT-284B2F/2020\|EPI_ISL_488525\|20 | .................... | .................... | .................... | .................... | .................... | .................... |
| E | 8551 | hCoV-19/England/NORT-2836E3/2020\|EPI_ISL_488526\|20 | .................... | .................... | .................... | .................... | .................... | .................... |
| E | 8552 | hCoV-19/England/NORT-2843DC/2020\|EPI_ISL_488527\|20 | .................... | .................... | .................... | .................... | .................... | .................... |
| E | 8553 | hCoV-19/England/NORT-282277/2020\|EPI_ISL_488528\|20 | .................... | .................... | .................... | .................... | .................... | .................... |
| E | 8554 | hCoV-19/England/NORT-2837FF/2020\|EPI_ISL_488529\|20 | .................... | .................... | .................... | .................... | .................... | .................... |
| E | 8555 | hCoV-19/England/NORT-295D34/2020\|EPI_ISL_488530\|20 | .................... | .................... | .................... | .................... | .................... | .................... |
| E | 8556 | hCoV-19/England/NORT-2958AC/2020\|EPI_ISL_488531\|20 | .................... | .................... | .................... | .................... | .................... | .................... |
| E | 8557 | hCoV-19/England/NORT-283BD5/2020\|EPI_ISL_488532\|20 | .................... | .................... | .................... | .................... | .................... | .................... |
| E | 8558 | hCoV-19/England/NORT-295648/2020\|EPI_ISL_488533\|20 | .................... | .................... | .................... | .................... | .................... | .................... |
| E | 8559 | hCoV-19/England/NORT-28224A/2020\|EPI_ISL_488536\|20 | .................... | .A.................. | .................... | .................... | .................... | .................... |
| E | 8560 | hCoV-19/England/NORT-2952AE/2020\|EPI_ISL_488545\|20 | .................... | .................... | .................... | .................... | .................... | .................... |
| E | 8561 | hCoV-19/England/NORT-283AF6/2020\|EPI_ISL_488547\|20 | .................... | .................... | .................... | .................... | .................... | .................... |
| E | 8562 | hCoV-19/England/NORT-283188/2020\|EPI_ISL_488548\|20 | .................... | .................... | .................... | .................... | .................... | .................... |
| E | 8563 | hCoV-19/England/NORT-282AE8/2020\|EPI_ISL_488549\|20 | .................... | .................... | .................... | .................... | .................... | .................... |
| E | 8564 | hCoV-19/England/NORT-294A2F/2020\|EPI_ISL_488550\|20 | .................... | .................... | .................... | .................... | .................... | .................... |
| E | 8565 | hCoV-19/England/NORT-295341/2020\|EPI_ISL_488551\|20 | .................... | .................... | .................... | .................... | .................... | .................... |
| E | 8566 | hCoV-19/England/NORT-294E50/2020\|EPI_ISL_488552\|20 | .................... | .................... | .................... | .................... | .................... | .................... |
| E | 8567 | hCoV-19/England/NORT-2843EB/2020\|EPI_ISL_488554\|20 | .................... | .................... | .................... | .................... | .................... | .................... |
| E | 8568 | hCoV-19/England/NORT-282AF7/2020\|EPI_ISL_488555\|20 | .................... | .................... | .................... | .................... | .................... | .................... |
| E | 8569 | hCoV-19/England/NORT-2831B5/2020\|EPI_ISL_488556\|20 | .................... | .................... | .................... | .................... | .................... | .................... |
| E | 8570 | hCoV-19/England/NORT-28408A/2020\|EPI_ISL_488557\|20 | .................... | .................... | .................... | .................... | .................... | .................... |
| E | 8571 | hCoV-19/England/NORT-2955A5/2020\|EPI_ISL_488558\|20 | .................... | .................... | .................... | .................... | .................... | .................... |

| | | | | | | | | |
|---|---|---|---|---|---|---|---|---|
| E | 8572 | hCoV-19/England/NORT-2835B9/2020\|EPI_ISL_488559\|20 | .................... | .................... | ..................... | .................... | .................... | .................... |
| E | 8573 | hCoV-19/England/NORT-28355F/2020\|EPI_ISL_488560\|20 | .................... | .................... | ..................... | .................... | .................... | .................... |
| E | 8574 | hCoV-19/England/NORT-283786/2020\|EPI_ISL_488561\|20 | .................... | .................... | ..................... | .................... | .................... | .................... |
| E | 8575 | hCoV-19/England/NORT-282E0A/2020\|EPI_ISL_488563\|20 | .................... | .................... | ..................... | .................... | .................... | .................... |
| E | 8576 | hCoV-19/England/NORT-2826F3/2020\|EPI_ISL_488564\|20 | .................... | .................... | ..................... | .................... | .................... | .................... |
| E | 8577 | hCoV-19/England/NORT-282B4F/2020\|EPI_ISL_488565\|20 | .................... | .................... | ..................... | .................... | .................... | .................... |
| E | 8578 | hCoV-19/England/NORT-2955C3/2020\|EPI_ISL_488566\|20 | .................... | .................... | ..................... | .................... | .................... | .................... |
| E | 8579 | hCoV-19/England/NORT-295CDD/2020\|EPI_ISL_488567\|20 | .................... | .................... | ..................... | .................... | .................... | .................... |
| E | 8580 | hCoV-19/England/NORT-283908/2020\|EPI_ISL_488568\|20 | .................... | .................... | ..................... | .................... | .................... | .................... |
| E | 8581 | hCoV-19/England/NORT-2830B8/2020\|EPI_ISL_488569\|20 | .................... | .................... | ..................... | .................... | .................... | .................... |
| E | 8582 | hCoV-19/England/NORT-295B1C/2020\|EPI_ISL_488570\|20 | .................... | .................... | ..................... | .................... | .................... | .................... |
| E | 8583 | hCoV-19/England/NORT-294C92/2020\|EPI_ISL_488571\|20 | .................... | .................... | ..................... | .................... | .................... | .................... |
| E | 8584 | hCoV-19/England/NORT-284390/2020\|EPI_ISL_488572\|20 | .................... | .................... | ..................... | .................... | .................... | .................... |
| E | 8585 | hCoV-19/England/NORT-2826D5/2020\|EPI_ISL_488573\|20 | .................... | .................... | ..................... | .................... | .................... | .................... |
| E | 8586 | hCoV-19/England/NORT-284DDE/2020\|EPI_ISL_488574\|20 | .................... | .................... | ..................... | .................... | .................... | .................... |
| E | 8587 | hCoV-19/England/NORT-2822C2/2020\|EPI_ISL_488575\|20 | .................... | .................... | ..................... | .................... | .................... | .................... |
| E | 8588 | hCoV-19/England/NORT-294AA7/2020\|EPI_ISL_488576\|20 | .................... | .................... | ..................... | .................... | .................... | .................... |
| E | 8589 | hCoV-19/England/NORT-2838DE/2020\|EPI_ISL_488577\|20 | .................... | .................... | ..................... | .................... | .................... | .................... |
| E | 8593 | hCoV-19/USA/FL-BPHL-0410/2020\|EPI_ISL_489819\|2020- | .................... | .................... | ..................... | .................... | .................... | .................... |
| E | 8594 | hCoV-19/USA/FL-BPHL-0414/2020\|EPI_ISL_489823\|2020- | .................... | .................... | ..................... | .................... | .................... | .................... |
| E | 8595 | hCoV-19/USA/FL-BPHL-0418/2020\|EPI_ISL_489827\|2020- | .................... | .................... | ..................... | .................... | .................... | .................... |
| E | 8596 | hCoV-19/USA/FL-BPHL-0419/2020\|EPI_ISL_489828\|2020- | .................... | .................... | ..................... | .................... | .................... | .................... |
| E | 8597 | hCoV-19/Spain/PV-HUB-51235489/2020\|EPI_ISL_489833\| | .................... | .................... | ..................... | .................... | .................... | .................... |
| E | 8598 | hCoV-19/Spain/PV-HUB-56592455/2020\|EPI_ISL_489834\| | .................... | .................... | ..................... | .................... | .................... | .................... |
| E | 8599 | hCoV-19/Spain/PV-HUB-50066191/2020\|EPI_ISL_489835\| | .................... | .................... | ..................... | .................... | .................... | .................... |
| E | 8600 | hCoV-19/Peru/LIM-INS-003/2020\|EPI_ISL_489836\|2020- | .................... | .................... | ..................... | .................... | .................... | .................... |
| E | 8601 | hCoV-19/Peru/LIM-INS-004/2020\|EPI_ISL_489837\|2020- | .................... | .................... | ..................... | .................... | .................... | .................... |
| E | 8602 | hCoV-19/Peru/LIM-INS-006/2020\|EPI_ISL_489838\|2020- | .................... | .................... | ..................... | .................... | .................... | .................... |
| E | 8603 | hCoV-19/Peru/HUC-INS-008/2020\|EPI_ISL_489839\|2020- | .................... | .................... | ..................... | .................... | .................... | .................... |
| E | 8604 | hCoV-19/USA/IA-GMF-08606/2020\|EPI_ISL_489896\|2020- | .................... | .A.................. | ..................... | .................... | .................... | .................... |
| E | 8605 | hCoV-19/USA/IA-GMF-08792/2020\|EPI_ISL_489897\|2020- | .................... | .A.................. | ..................... | .................... | .................... | .................... |
| E | 8606 | hCoV-19/USA/IA-GMF-08809/2020\|EPI_ISL_489898\|2020- | .................... | .A.................. | ..................... | .................... | .................... | .................... |
| E | 8607 | hCoV-19/Wales/PHWC-165DA7/2020\|EPI_ISL_490800\|2020 | .................... | .................... | ..................... | .................... | .................... | .................... |
| E | 8608 | hCoV-19/Wales/PHWC-165DC5/2020\|EPI_ISL_490802\|2020 | .................... | .................... | ..................... | .................... | .................... | .................... |
| E | 8609 | hCoV-19/Wales/PHWC-165E0E/2020\|EPI_ISL_490805\|2020 | .................... | .................... | ..................... | .................... | .................... | .................... |
| E | 8610 | hCoV-19/Wales/PHWC-165E2C/2020\|EPI_ISL_490806\|2020 | .................... | .................... | ..................... | .................... | .................... | .................... |
| E | 8611 | hCoV-19/Wales/PHWC-165E68/2020\|EPI_ISL_490807\|2020 | .................... | .................... | ..................... | .................... | .................... | .................... |
| E | 8612 | hCoV-19/Wales/PHWC-165EC2/2020\|EPI_ISL_490812\|2020 | .................... | .................... | ..................... | .................... | .................... | .................... |
| E | 8613 | hCoV-19/Wales/PHWC-165EFF/2020\|EPI_ISL_490815\|2020 | .................... | .................... | ..................... | .................... | .................... | .................... |
| E | 8614 | hCoV-19/Wales/PHWC-165F56/2020\|EPI_ISL_490818\|2020 | .................... | .................... | ..................... | .................... | .................... | .................... |
| E | 8615 | hCoV-19/Wales/PHWC-165F92/2020\|EPI_ISL_490820\|2020 | .................... | .................... | ..................... | .................... | .................... | .................... |
| E | 8616 | hCoV-19/Wales/PHWC-165FFC/2020\|EPI_ISL_490825\|2020 | .................... | .................... | ..................... | .................... | .................... | .................... |
| E | 8617 | hCoV-19/Wales/PHWC-166025/2020\|EPI_ISL_490827\|2020 | .................... | .................... | ..................... | .................... | .................... | .................... |
| E | 8618 | hCoV-19/Wales/PHWC-166061/2020\|EPI_ISL_490829\|2020 | .................... | .................... | ..................... | .................... | .................... | .................... |
| E | 8619 | hCoV-19/Wales/PHWC-166070/2020\|EPI_ISL_490830\|2020 | .................... | .................... | ..................... | .................... | .................... | .................... |
| E | 8620 | hCoV-19/Wales/PHWC-1660AD/2020\|EPI_ISL_490833\|2020 | .................... | .................... | ..................... | .................... | .................... | .................... |
| E | 8621 | hCoV-19/Wales/PHWC-1660DA/2020\|EPI_ISL_490835\|2020 | .................... | .................... | ..................... | .................... | .................... | .................... |
| E | 8622 | hCoV-19/Wales/PHWC-166113/2020\|EPI_ISL_490838\|2020 | .................... | .................... | ..................... | .................... | .................... | .................... |
| E | 8623 | hCoV-19/Wales/PHWC-166122/2020\|EPI_ISL_490839\|2020 | .................... | .................... | ..................... | .................... | .................... | .................... |
| E | 8624 | hCoV-19/Wales/PHWC-166140/2020\|EPI_ISL_490840\|2020 | .................... | .................... | ..................... | .................... | .................... | .................... |
| E | 8625 | hCoV-19/Wales/PHWC-16619B/2020\|EPI_ISL_490845\|2020 | .................... | .................... | ..................... | .................... | .................... | .................... |
| E | 8626 | hCoV-19/Wales/PHWC-1661D7/2020\|EPI_ISL_490848\|2020 | .................... | .................... | ..................... | .................... | .................... | .................... |
| E | 8627 | hCoV-19/Wales/PHWC-1661F5/2020\|EPI_ISL_490850\|2020 | .................... | .................... | ..................... | .................... | .................... | .................... |
| E | 8628 | hCoV-19/Wales/PHWC-166201/2020\|EPI_ISL_490851\|2020 | .................... | .................... | ..................... | .................... | .................... | .................... |
| E | 8629 | hCoV-19/Wales/PHWC-166210/2020\|EPI_ISL_490852\|2020 | .................... | .................... | ..................... | .................... | .................... | .................... |
| E | 8630 | hCoV-19/Wales/PHWC-16624D/2020\|EPI_ISL_490855\|2020 | .................... | .................... | ..................... | .................... | .................... | .................... |
| E | 8631 | hCoV-19/Wales/PHWC-166298/2020\|EPI_ISL_490857\|2020 | .................... | .................... | ..................... | .................... | .................... | .................... |
| E | 8632 | hCoV-19/Wales/PHWC-1662B6/2020\|EPI_ISL_490859\|2020 | .................... | .................... | ..................... | .................... | .................... | .................... |
| E | 8633 | hCoV-19/USA/UT-03278/2020\|EPI_ISL_493011\|2020-05-0 | .................... | .................... | ..................... | .................... | .................... | .................... |
| E | 8634 | hCoV-19/USA/UT-03279/2020\|EPI_ISL_493012\|2020-05-0 | .................... | .................... | ..................... | .................... | .................... | .................... |
| E | 8635 | hCoV-19/USA/UT-03282/2020\|EPI_ISL_493013\|2020-05-0 | .................... | .................... | ..................... | .................... | .................... | .................... |
| E | 8636 | hCoV-19/USA/UT-03283/2020\|EPI_ISL_493014\|2020-05-0 | .................... | .................... | ..................... | .................... | .................... | .................... |
| E | 8637 | hCoV-19/USA/UT-03288/2020\|EPI_ISL_493015\|2020-05-0 | .................... | .................... | ..................... | .................... | .................... | .................... |
| E | 8638 | hCoV-19/USA/UT-03290/2020\|EPI_ISL_493016\|2020-05-0 | .................... | .................... | ..................... | .................... | .................... | .................... |
| E | 8639 | hCoV-19/USA/UT-03293/2020\|EPI_ISL_493017\|2020-05-0 | .................... | .................... | ..................... | .................... | .................... | .................... |
| E | 8640 | hCoV-19/USA/UT-03296/2020\|EPI_ISL_493018\|2020-05-0 | .................... | .................... | ..................... | .................... | .................... | .................... |
| E | 8641 | hCoV-19/USA/UT-03302/2020\|EPI_ISL_493019\|2020-05-0 | .................... | .................... | ..................... | .................... | .................... | .................... |
| E | 8642 | hCoV-19/USA/UT-03305/2020\|EPI_ISL_493020\|2020-05-0 | .................... | .................... | ..................... | .................... | .................... | .................... |
| E | 8643 | hCoV-19/USA/UT-03306/2020\|EPI_ISL_493021\|2020-05-0 | .................... | .................... | ..................... | .................... | .................... | .................... |
| E | 8644 | hCoV-19/USA/UT-04172/2020\|EPI_ISL_493050\|2020-05-1 | .................... | .................... | ..................... | .................... | .................... | .................... |
| E | 8645 | hCoV-19/USA/UT-04176/2020\|EPI_ISL_493051\|2020-05-1 | .................... | .................... | ..................... | .................... | .................... | .................... |
| E | 8646 | hCoV-19/USA/WY-WYPHL-20006826/2020\|EPI_ISL_493061\| | .................... | .................... | ..................... | .................... | .................... | .................... |
| E | 8647 | hCoV-19/USA/WY-WYPHL-20006987/2020\|EPI_ISL_493062\| | .................... | .................... | ..................... | .................... | .................... | .................... |
| E | 8648 | hCoV-19/USA/WY-WYPHL-20007007/2020\|EPI_ISL_493063\| | .................... | .................... | ..................... | .................... | .................... | .................... |
| E | 8649 | hCoV-19/USA/WY-WYPHL-20007405/2020\|EPI_ISL_493064\| | .................... | .................... | ..................... | .................... | .................... | .................... |
| E | 8650 | hCoV-19/USA/MO-WUSTL031/2020\|EPI_ISL_493069\|2020-0 | .................... | .................... | ..................... | .................... | .................... | .................... |
| E | 8651 | hCoV-19/USA/MO-WUSTL034/2020\|EPI_ISL_493070\|2020-0 | .................... | .................... | ..................... | .................... | .................... | .................... |
| E | 8652 | hCoV-19/USA/MO-WUSTL036/2020\|EPI_ISL_493071\|2020-0 | .................... | .................... | ..................... | .................... | .................... | .................... |
| E | 8653 | hCoV-19/USA/MO-WUSTL037/2020\|EPI_ISL_493072\|2020-0 | .................... | .................... | ..................... | .................... | .................... | .................... |
| E | 8654 | hCoV-19/USA/MO-WUSTL042/2020\|EPI_ISL_493073\|2020-0 | .................... | .................... | ..................... | .................... | .................... | .................... |

| | | | | | | | | |
|---|---|---|---|---|---|---|---|---|
| E | 8655 | hCoV-19/USA/MO-WUSTL043/2020\|EPI_ISL_493074\|2020-0 | ···················· | ···················· | ····················· | ···················· | ···················· | ···················· |
| E | 8656 | hCoV-19/USA/MO-WUSTL046/2020\|EPI_ISL_493075\|2020-0 | ···················· | ···················· | ····················· | ···················· | ···················· | ···················· |
| E | 8657 | hCoV-19/USA/MO-WUSTL047/2020\|EPI_ISL_493076\|2020-0 | ···················· | ···················· | ····················· | ···················· | ···················· | ···················· |
| E | 8658 | hCoV-19/USA/MO-WUSTL050/2020\|EPI_ISL_493077\|2020-0 | ···················· | ···················· | ····················· | ···················· | ···················· | ···················· |
| E | 8659 | hCoV-19/USA/MO-WUSTL060/2020\|EPI_ISL_493079\|2020-0 | ···················· | ···················· | ····················· | ···················· | ···················· | ···················· |
| E | 8660 | hCoV-19/USA/MO-WUSTL061/2020\|EPI_ISL_493080\|2020-0 | ···················· | ···················· | ····················· | ···················· | ···················· | ···················· |
| E | 8661 | hCoV-19/USA/MO-WUSTL062/2020\|EPI_ISL_493081\|2020-0 | ···················· | ···················· | ····················· | ···················· | ···················· | ···················· |
| E | 8662 | hCoV-19/USA/MO-WUSTL063/2020\|EPI_ISL_493082\|2020-0 | ···················· | ···················· | ····················· | ···················· | ···················· | ···················· |
| E | 8663 | hCoV-19/USA/MO-WUSTL064/2020\|EPI_ISL_493083\|2020-0 | ···················· | ···················· | ····················· | ···················· | ···················· | ···················· |
| E | 8664 | hCoV-19/USA/MO-WUSTL067/2020\|EPI_ISL_493084\|2020-0 | ···················· | ···················· | ····················· | ···················· | ···················· | ···················· |
| E | 8665 | hCoV-19/USA/MO-WUSTL068/2020\|EPI_ISL_493085\|2020-0 | ···················· | ···G················ | ····················· | ···················· | ···················· | ···················· |
| E | 8666 | hCoV-19/USA/UT-02448/2020\|EPI_ISL_493089\|2020-04-2 | ···················· | ···················· | ····················· | ···················· | ···················· | ···················· |
| E | 8667 | hCoV-19/USA/UT-02449/2020\|EPI_ISL_493090\|2020-04-2 | ···················· | ···················· | ····················· | ···················· | ···················· | ···················· |
| E | 8668 | hCoV-19/USA/UT-02455/2020\|EPI_ISL_493096\|2020-04-2 | ···················· | ···················· | ····················· | ···················· | ···················· | ···················· |
| E | 8669 | hCoV-19/USA/UT-02510/2020\|EPI_ISL_493098\|2020-04-2 | ···················· | ···················· | ····················· | ···················· | ···················· | ···················· |
| E | 8670 | hCoV-19/USA/UT-02511/2020\|EPI_ISL_493099\|2020-04-2 | ···················· | ···················· | ····················· | ···················· | ···················· | ···················· |
| E | 8671 | hCoV-19/Wales/PHWC-168BE6/2020\|EPI_ISL_494344\|2020 | ···················· | ···A················ | ····················· | ···················· | ···················· | ···················· |
| E | 8672 | hCoV-19/Wales/PHWC-168CB6/2020\|EPI_ISL_494351\|2020 | ···················· | ···················· | ····················· | ···················· | ···················· | ···················· |
| E | 8673 | hCoV-19/USA/CA-ALSR-1123/2020\|EPI_ISL_494375\|2020- | ···················· | ···················· | ····················· | ···················· | ···················· | ···················· |
| E | 8674 | hCoV-19/USA/CA-ALSR-1124/2020\|EPI_ISL_494376\|2020- | ···················· | ·A·················· | ····················· | ···················· | ···················· | ···················· |
| E | 8675 | hCoV-19/USA/CA-ALSR-1125/2020\|EPI_ISL_494377\|2020- | ···················· | ···················· | ····················· | ···················· | ···················· | ···················· |
| E | 8676 | hCoV-19/USA/CA-ALSR-1126/2020\|EPI_ISL_494378\|2020- | ···················· | ···················· | ····················· | ···················· | ···················· | ···················· |
| E | 8677 | hCoV-19/USA/CA-ALSR-1127/2020\|EPI_ISL_494379\|2020- | ···················· | ···················· | ····················· | ···················· | ···················· | ···················· |
| E | 8678 | hCoV-19/USA/CA-ALSR-1129/2020\|EPI_ISL_494380\|2020- | ···················· | ···················· | ····················· | ···················· | ···················· | ···················· |
| E | 8679 | hCoV-19/USA/CA-ALSR-1130/2020\|EPI_ISL_494381\|2020- | ···················· | ···················· | ····················· | ···················· | ···················· | ···················· |
| E | 8680 | hCoV-19/USA/CA-ALSR-1132/2020\|EPI_ISL_494382\|2020- | ···················· | ·A·················· | ····················· | ···················· | ···················· | ···················· |
| E | 8681 | hCoV-19/USA/CA-ALSR-1135/2020\|EPI_ISL_494383\|2020- | ···················· | ···················· | ····················· | ···················· | ···················· | ···················· |
| E | 8682 | hCoV-19/USA/CA-ALSR-1138/2020\|EPI_ISL_494384\|2020- | ···················· | ···················· | ····················· | ···················· | ···················· | ···················· |
| E | 8683 | hCoV-19/USA/CA-ALSR-1140/2020\|EPI_ISL_494385\|2020- | ···················· | ···················· | ····················· | ···················· | ···················· | ···················· |
| E | 8684 | hCoV-19/USA/CA-ALSR-1141/2020\|EPI_ISL_494386\|2020- | ···················· | ·A·················· | ····················· | ···················· | ···················· | ···················· |
| E | 8685 | hCoV-19/USA/CA-ALSR-1142/2020\|EPI_ISL_494387\|2020- | ···················· | ···················· | ····················· | ···················· | ···················· | ···················· |
| E | 8686 | hCoV-19/USA/CA-ALSR-1143/2020\|EPI_ISL_494388\|2020- | ···················· | ·A·················· | ····················· | ···················· | ···················· | ···················· |
| E | 8687 | hCoV-19/USA/CA-ALSR-1144/2020\|EPI_ISL_494389\|2020- | ···················· | ·A·················· | ····················· | ···················· | ···················· | ···················· |
| E | 8688 | hCoV-19/USA/CA-ALSR-1145/2020\|EPI_ISL_494390\|2020- | ···················· | ···················· | ····················· | ···················· | ···················· | ···················· |
| E | 8689 | hCoV-19/USA/CA-ALSR-1146/2020\|EPI_ISL_494391\|2020- | ···················· | ···················· | ····················· | ···················· | ···················· | ···················· |
| E | 8690 | hCoV-19/USA/CA-ALSR-1147/2020\|EPI_ISL_494392\|2020- | ···················· | ···················· | ····················· | ···················· | ···················· | ···················· |
| E | 8691 | hCoV-19/USA/CA-ALSR-1148/2020\|EPI_ISL_494393\|2020- | ···················· | ·A·················· | ····················· | ···················· | ···················· | ···················· |
| E | 8692 | hCoV-19/USA/CA-ALSR-1151/2020\|EPI_ISL_494394\|2020- | ···················· | ···················· | ····················· | ···················· | ···················· | ···················· |
| E | 8693 | hCoV-19/USA/CA-ALSR-1153/2020\|EPI_ISL_494396\|2020- | ···················· | ·A·················· | ·········T··········· | ···················· | ···················· | ···················· |
| E | 8694 | hCoV-19/USA/CA-ALSR-1154/2020\|EPI_ISL_494397\|2020- | ···················· | ·A·················· | ····················· | ···················· | ···················· | ···················· |
| E | 8695 | hCoV-19/USA/CA-ALSR-1155/2020\|EPI_ISL_494398\|2020- | ···················· | ···················· | ····················· | ···················· | ···················· | ···················· |
| E | 8696 | hCoV-19/USA/CA-ALSR-1156/2020\|EPI_ISL_494399\|2020- | ···················· | ·A·················· | ····················· | ···················· | ···················· | ···················· |
| E | 8697 | hCoV-19/USA/WA-S1223/2020\|EPI_ISL_495673\|2020-05-0 | ···················· | ···················· | ····················· | ···················· | ···················· | ···················· |
| E | 8698 | hCoV-19/USA/WA-S1224/2020\|EPI_ISL_495674\|2020-05-0 | ···················· | ···················· | ····················· | ···················· | ···················· | ···················· |
| E | 8699 | hCoV-19/USA/WA-S1226/2020\|EPI_ISL_495676\|2020-05-0 | ···················· | ···················· | ····················· | ···················· | ···················· | ···················· |
| E | 8700 | hCoV-19/USA/WA-S1227/2020\|EPI_ISL_495677\|2020-05-0 | ···················· | ···················· | ····················· | ···················· | ···················· | ···················· |
| E | 8701 | hCoV-19/USA/WA-S1230/2020\|EPI_ISL_495680\|2020-05-0 | ···················· | ···················· | ····················· | ···················· | ···················· | ···················· |
| E | 8702 | hCoV-19/USA/WA-S1231/2020\|EPI_ISL_495681\|2020-05-0 | ···················· | ···················· | ····················· | ···················· | ···················· | ···················· |
| E | 8703 | hCoV-19/USA/WA-S1232/2020\|EPI_ISL_495682\|2020-05-0 | ···················· | ···················· | ····················· | ···················· | ···················· | ···················· |
| E | 8704 | hCoV-19/USA/WA-S1233/2020\|EPI_ISL_495683\|2020-05-0 | ···················· | ···················· | ····················· | ···················· | ···················· | ···················· |
| E | 8745 | hCoV-19/USA/OR-PROV-026/2020\|EPI_ISL_482310\|2020-0 | ···················· | ···················· | ····················· | ···················· | ···················· | ···················· |
| E | 8746 | hCoV-19/USA/OR-PROV-027/2020\|EPI_ISL_482311\|2020-0 | ···················· | ···················· | ····················· | ···················· | ···················· | ···················· |
| E | 8747 | hCoV-19/USA/OR-PROV-028/2020\|EPI_ISL_482312\|2020-0 | ···················· | ···················· | ····················· | ···················· | ···················· | ···················· |
| E | 8748 | hCoV-19/USA/OR-PROV-030/2020\|EPI_ISL_482313\|2020-0 | ···················· | ···················· | ····················· | ···················· | ···················· | ···················· |
| E | 8749 | hCoV-19/USA/OR-PROV-031/2020\|EPI_ISL_482314\|2020-0 | ···················· | ···················· | ····················· | ···················· | ···················· | ···················· |
| E | 8750 | hCoV-19/USA/OR-PROV-032/2020\|EPI_ISL_482315\|2020-0 | ···················· | ···················· | ····················· | ···················· | ···················· | ···················· |
| E | 8751 | hCoV-19/USA/OR-PROV-033/2020\|EPI_ISL_482316\|2020-0 | ···················· | ···················· | ····················· | ···················· | ···················· | ···················· |
| E | 8752 | hCoV-19/USA/OR-PROV-035/2020\|EPI_ISL_482317\|2020-0 | ···················· | ···················· | ····················· | ···················· | ···················· | ···················· |
| E | 8753 | hCoV-19/USA/OR-PROV-036/2020\|EPI_ISL_482318\|2020-0 | ···················· | ···················· | ····················· | ···················· | ···················· | ···················· |
| E | 8754 | hCoV-19/USA/OR-PROV-037/2020\|EPI_ISL_482319\|2020-0 | ···················· | ···················· | ····················· | ···················· | ···················· | ···················· |
| E | 8755 | hCoV-19/USA/OR-PROV-038/2020\|EPI_ISL_482320\|2020-0 | ···················· | ···················· | ····················· | ···················· | ···················· | ···················· |
| E | 8756 | hCoV-19/USA/OR-PROV-039/2020\|EPI_ISL_482321\|2020-0 | ···················· | ···················· | ····················· | ···················· | ···················· | ···················· |
| E | 8757 | hCoV-19/USA/OR-PROV-040/2020\|EPI_ISL_482322\|2020-0 | ···················· | ···················· | ····················· | ···················· | ···················· | ···················· |
| E | 8758 | hCoV-19/USA/OR-PROV-041/2020\|EPI_ISL_482323\|2020-0 | ···················· | ···················· | ····················· | ···················· | ···················· | ···················· |
| E | 8759 | hCoV-19/USA/OR-PROV-042/2020\|EPI_ISL_482324\|2020-0 | ···················· | ···················· | ····················· | ···················· | ···················· | ···················· |
| E | 8760 | hCoV-19/USA/OR-PROV-043/2020\|EPI_ISL_482325\|2020-0 | ···················· | ···················· | ····················· | ···················· | ···················· | ···················· |
| E | 8761 | hCoV-19/USA/OR-PROV-044/2020\|EPI_ISL_482326\|2020-0 | ···················· | ···················· | ····················· | ···················· | ···················· | ···················· |
| E | 8762 | hCoV-19/USA/OR-PROV-045/2020\|EPI_ISL_482327\|2020-0 | ···················· | ···················· | ····················· | ···················· | ···················· | ···················· |
| E | 8763 | hCoV-19/USA/OR-PROV-046/2020\|EPI_ISL_482328\|2020-0 | ···················· | ···················· | ····················· | ···················· | ···················· | ···················· |
| E | 8764 | hCoV-19/USA/OR-PROV-048/2020\|EPI_ISL_482329\|2020-0 | ···················· | ···················· | ····················· | ···················· | ···················· | ···················· |
| E | 8765 | hCoV-19/USA/OR-PROV-050/2020\|EPI_ISL_482330\|2020-0 | ···················· | ···················· | ····················· | ···················· | ···················· | ···················· |
| E | 8766 | hCoV-19/USA/OR-PROV-051/2020\|EPI_ISL_482331\|2020-0 | ···················· | ···················· | ····················· | ···················· | ···················· | ···················· |
| E | 8767 | hCoV-19/USA/OR-PROV-052/2020\|EPI_ISL_482332\|2020-0 | ···················· | ···················· | ····················· | ···················· | ···················· | ···················· |
| E | 8768 | hCoV-19/USA/OR-PROV-053/2020\|EPI_ISL_482333\|2020-0 | ···················· | ···················· | ····················· | ···················· | ···················· | ···················· |
| E | 8769 | hCoV-19/USA/OR-PROV-054/2020\|EPI_ISL_482334\|2020-0 | ···················· | ···················· | ····················· | ···················· | ···················· | ···················· |
| E | 8770 | hCoV-19/USA/OR-PROV-055/2020\|EPI_ISL_482335\|2020-0 | ···················· | ···················· | ····················· | ···················· | ···················· | ···················· |
| E | 8771 | hCoV-19/USA/OR-PROV-056/2020\|EPI_ISL_482336\|2020-0 | ···················· | ···················· | ····················· | ···················· | ···················· | ···················· |
| E | 8772 | hCoV-19/USA/OR-PROV-058/2020\|EPI_ISL_482338\|2020-0 | ···················· | ···················· | ····················· | ···················· | ···················· | ···················· |
| E | 8773 | hCoV-19/USA/OR-PROV-059/2020\|EPI_ISL_482339\|2020-0 | ···················· | ···················· | ····················· | ···················· | ···················· | ···················· |
| E | 8774 | hCoV-19/USA/OR-PROV-060/2020\|EPI_ISL_482340\|2020-0 | ···················· | ···················· | ····················· | ···················· | ···················· | ···················· |

| | | | | | | | | |
|---|---|---|---|---|---|---|---|---|
| E | 8775 | hCoV-19/USA/OR-PROV-061/2020\|EPI_ISL_482341\|2020-0 | ···················· | ···················· | ···················· | ···················· | ···················· | ···················· |
| E | 8776 | hCoV-19/USA/OR-PROV-062/2020\|EPI_ISL_482342\|2020-0 | ···················· | ···················· | ···················· | ···················· | ···················· | ···················· |
| E | 8777 | hCoV-19/USA/OR-PROV-064/2020\|EPI_ISL_482343\|2020-0 | ···················· | ···················· | ···················· | ···················· | ···················· | ···················· |
| E | 8778 | hCoV-19/USA/OR-PROV-065/2020\|EPI_ISL_482344\|2020-0 | ···················· | ···················· | ···················· | ···················· | ···················· | ···················· |
| E | 8779 | hCoV-19/USA/OR-PROV-070/2020\|EPI_ISL_482347\|2020-0 | ···················· | ···················· | ···················· | ···················· | ···················· | ···················· |
| E | 8780 | hCoV-19/USA/OR-PROV-073/2020\|EPI_ISL_482348\|2020-0 | ···················· | ···················· | ···················· | ···················· | ···················· | ···················· |
| E | 8781 | hCoV-19/USA/OR-PROV-075/2020\|EPI_ISL_482350\|2020-0 | ···················· | ·A·················· | ···················· | ···················· | ···················· | ···················· |
| E | 8782 | hCoV-19/USA/OR-PROV-078/2020\|EPI_ISL_482351\|2020-0 | ···················· | ···················· | ···················· | ···················· | ···················· | ···················· |
| E | 8783 | hCoV-19/USA/OR-PROV-093/2020\|EPI_ISL_482364\|2020-0 | ···················· | ···················· | ···················· | ···················· | ···················· | ···················· |
| E | 8784 | hCoV-19/USA/OR-PROV-094/2020\|EPI_ISL_482365\|2020-0 | ···················· | ···················· | ···················· | ···················· | ···················· | ···················· |
| E | 8785 | hCoV-19/USA/OR-PROV-095/2020\|EPI_ISL_482366\|2020-0 | ···················· | ···················· | ···················· | ···················· | ···················· | ···················· |
| E | 8786 | hCoV-19/USA/OR-PROV-096/2020\|EPI_ISL_482367\|2020-0 | ···················· | ···················· | ···················· | ···················· | ···················· | ···················· |
| E | 8787 | hCoV-19/USA/OR-PROV-097/2020\|EPI_ISL_482368\|2020-0 | ···················· | ·A·················· | ···················· | ···················· | ···················· | ···················· |
| E | 8788 | hCoV-19/USA/OR-PROV-098/2020\|EPI_ISL_482369\|2020-0 | ···················· | ···················· | ···················· | ···················· | ···················· | ···················· |
| E | 8789 | hCoV-19/USA/OR-PROV-100/2020\|EPI_ISL_482370\|2020-0 | ···················· | ···················· | ···················· | ···················· | ···················· | ···················· |
| E | 8790 | hCoV-19/USA/OR-PROV-101/2020\|EPI_ISL_482371\|2020-0 | ···················· | ···················· | ···················· | ···················· | ···················· | ···················· |
| E | 8791 | hCoV-19/USA/OR-PROV-102/2020\|EPI_ISL_482372\|2020-0 | ···················· | ···················· | ···················· | ···················· | ···················· | ···················· |
| E | 8792 | hCoV-19/USA/OR-PROV-103/2020\|EPI_ISL_482373\|2020-0 | ···················· | ···················· | ···················· | ···················· | ···················· | ···················· |
| E | 8793 | hCoV-19/USA/OR-PROV-106/2020\|EPI_ISL_482374\|2020-0 | ···················· | ···················· | ···················· | ···················· | ···················· | ···················· |
| E | 8794 | hCoV-19/USA/OR-PROV-107/2020\|EPI_ISL_482375\|2020-0 | ···················· | ···················· | ···················· | ···················· | ···················· | ···················· |
| E | 8795 | hCoV-19/USA/OR-PROV-108/2020\|EPI_ISL_482376\|2020-0 | ···················· | ···················· | ···················· | ···················· | ···················· | ···················· |
| E | 8796 | hCoV-19/USA/OR-PROV-109/2020\|EPI_ISL_482377\|2020-0 | ···················· | ···················· | ···················· | ···················· | ···················· | ···················· |
| E | 8797 | hCoV-19/USA/OR-PROV-110/2020\|EPI_ISL_482378\|2020-0 | ···················· | ···················· | ···················· | ···················· | ···················· | ···················· |
| E | 8798 | hCoV-19/USA/OR-PROV-112/2020\|EPI_ISL_482379\|2020-0 | ···················· | ···················· | ···················· | ···················· | ···················· | ···················· |
| E | 8799 | hCoV-19/USA/OR-PROV-113/2020\|EPI_ISL_482380\|2020-0 | ···················· | ···················· | ···················· | ···················· | ···················· | ···················· |
| E | 8800 | hCoV-19/USA/OR-PROV-115/2020\|EPI_ISL_482381\|2020-0 | ···················· | ···················· | ···················· | ···················· | ···················· | ···················· |
| E | 8801 | hCoV-19/USA/OR-PROV-117/2020\|EPI_ISL_482382\|2020-0 | ···················· | ···················· | ···················· | ···················· | ···················· | ···················· |
| E | 8802 | hCoV-19/USA/OR-PROV-118/2020\|EPI_ISL_482383\|2020-0 | ···················· | ···················· | ···················· | ···················· | ···················· | ···················· |
| E | 8803 | hCoV-19/USA/OR-PROV-120/2020\|EPI_ISL_482384\|2020-0 | ···················· | ···················· | ···················· | ···················· | ···················· | ···················· |
| E | 8804 | hCoV-19/USA/OR-PROV-121/2020\|EPI_ISL_482385\|2020-0 | ···················· | ···················· | ···················· | ···················· | ···················· | ···················· |
| E | 8805 | hCoV-19/USA/OR-PROV-122/2020\|EPI_ISL_482386\|2020-0 | ···················· | ···················· | ···················· | ···················· | ···················· | ···················· |
| E | 8806 | hCoV-19/USA/OR-PROV-123/2020\|EPI_ISL_482387\|2020-0 | ···················· | ···················· | ···················· | ···················· | ···················· | ···················· |
| E | 8807 | hCoV-19/USA/OR-PROV-124/2020\|EPI_ISL_482388\|2020-0 | ···················· | ···················· | ···················· | ···················· | ···················· | ···················· |
| E | 8808 | hCoV-19/USA/OR-PROV-125/2020\|EPI_ISL_482389\|2020-0 | ···················· | ···················· | ···················· | ···················· | ···················· | ···················· |
| E | 8809 | hCoV-19/USA/OR-PROV-126/2020\|EPI_ISL_482390\|2020-0 | ···················· | ···················· | ···················· | ···················· | ···················· | ···················· |
| E | 8810 | hCoV-19/USA/OR-PROV-128/2020\|EPI_ISL_482392\|2020-0 | ···················· | ···················· | ···················· | ···················· | ···················· | ···················· |
| E | 8811 | hCoV-19/USA/OR-PROV-129/2020\|EPI_ISL_482393\|2020-0 | ···················· | ···················· | ···················· | ···················· | ···················· | ···················· |
| E | 8812 | hCoV-19/USA/OR-PROV-130/2020\|EPI_ISL_482394\|2020-0 | ···················· | ···················· | ···················· | ···················· | ···················· | ···················· |
| E | 8813 | hCoV-19/USA/OR-PROV-132/2020\|EPI_ISL_482395\|2020-0 | ···················· | ···················· | ···················· | ···················· | ···················· | ···················· |
| E | 8814 | hCoV-19/Bangladesh/BCSIR-NILMRC-195/2020\|EPI_ISL_4 | ···················· | ···················· | ···················· | ···················· | ···················· | ···················· |
| E | 8815 | hCoV-19/Bangladesh/BCSIR-NILMRC-198/2020\|EPI_ISL_4 | ···················· | ···················· | ···················· | ···················· | ···················· | ···················· |
| E | 8816 | hCoV-19/Bangladesh/BCSIR-NILMRC-199/2020\|EPI_ISL_4 | ···················· | ···················· | ···················· | ···················· | ···················· | ···················· |
| E | 8817 | hCoV-19/Bangladesh/BCSIR-NILMRC-201/2020\|EPI_ISL_4 | ···················· | ···················· | ···················· | ···················· | ···················· | ···················· |
| E | 8818 | hCoV-19/Bangladesh/BCSIR-NILMRC-211/2020\|EPI_ISL_4 | ···················· | ···················· | ···················· | ···················· | ···················· | ···················· |
| E | 8819 | hCoV-19/Bangladesh/BCSIR-NILMRC-212/2020\|EPI_ISL_4 | ···················· | ···················· | ···················· | ···················· | ···················· | ···················· |
| E | 8820 | hCoV-19/Switzerland/ZH-ETHZ-150000plus/2020\|EPI_IS | ···················· | ···················· | ···················· | ···················· | ···················· | ···················· |
| E | 8821 | hCoV-19/Switzerland/ZH-ETHZ-150001plus/2020\|EPI_IS | ···················· | ···················· | ···················· | ···················· | ···················· | ···················· |
| E | 8822 | hCoV-19/Switzerland/BE-ETHZ-150006plus/2020\|EPI_IS | ···················· | ···················· | ···················· | ···················· | ···················· | ···················· |
| E | 8823 | hCoV-19/Switzerland/BE-ETHZ-150012plus/2020\|EPI_IS | ···················· | ···················· | ···················· | ···················· | ···················· | ···················· |
| E | 8824 | hCoV-19/Switzerland/BL-ETHZ-150014plus/2020\|EPI_IS | ···················· | ···················· | ···················· | ···················· | ···················· | ···················· |
| E | 8825 | hCoV-19/Switzerland/BE-ETHZ-150015plus/2020\|EPI_IS | ···················· | ···················· | ···················· | ···················· | ···················· | ···················· |
| E | 8826 | hCoV-19/Switzerland/VD-ETHZ-150017plus/2020\|EPI_IS | ···················· | ···················· | ···················· | ···················· | ···················· | ···················· |
| E | 8827 | hCoV-19/Switzerland/ZG-ETHZ-150018plus/2020\|EPI_IS | ···················· | ···················· | ···················· | ···················· | ···················· | ···················· |
| E | 8828 | hCoV-19/Switzerland/BE-ETHZ-150027plus/2020\|EPI_IS | ···················· | ···················· | ···················· | ···················· | ···················· | ···················· |
| E | 8829 | hCoV-19/Switzerland/ZH-ETHZ-150029plus/2020\|EPI_IS | ···················· | ···················· | ···················· | ···················· | ···················· | ···················· |
| E | 8830 | hCoV-19/Switzerland/ZH-ETHZ-150034plus/2020\|EPI_IS | ···················· | ···················· | ···················· | ···················· | ···················· | ···················· |
| E | 8831 | hCoV-19/Switzerland/GE-ETHZ-150035plus/2020\|EPI_IS | ···················· | ·R·················· | ···················· | ···················· | ···················· | ···················· |
| E | 8832 | hCoV-19/Switzerland/VD-ETHZ-150036plus/2020\|EPI_IS | ···················· | ···················· | ···················· | ···················· | ···················· | ···················· |
| E | 8833 | hCoV-19/Switzerland/ZH-ETHZ-150040plus/2020\|EPI_IS | ···················· | ···················· | ···················· | ···················· | ···················· | ···················· |
| E | 8834 | hCoV-19/Switzerland/ZH-ETHZ-150041plus/2020\|EPI_IS | ···················· | ···················· | ···················· | ···················· | ···················· | ···················· |
| E | 8835 | hCoV-19/Switzerland/ZH-ETHZ-722plus/2020\|EPI_ISL_4 | ···················· | ···················· | ···················· | ···················· | ···················· | ···················· |
| E | 8836 | hCoV-19/Switzerland/ZH-ETHZ-381/2020\|EPI_ISL_48367 | ···················· | ···················· | ···················· | ···················· | ···················· | ······R············· |
| E | 8838 | hCoV-19/Switzerland/ZH-ETHZ-720plus/2020\|EPI_ISL_4 | ···················· | ···················· | ···················· | ···················· | ···················· | ···················· |
| E | 8839 | hCoV-19/Switzerland/ZH-ETHZ-838/2020\|EPI_ISL_48368 | ···················· | ···················· | ···················· | ···················· | ···················· | ···················· |
| E | 8840 | hCoV-19/Bangladesh/BCSIR-NILMRC-186/2020\|EPI_ISL_4 | ···················· | ·A·················· | ···················· | ···················· | ···················· | ··A················· |
| E | 8841 | hCoV-19/Bangladesh/BCSIR-NILMRC-190/2020\|EPI_ISL_4 | ···················· | ···················· | ···················· | ···················· | ···················· | ···················· |
| E | 8842 | hCoV-19/Bangladesh/BCSIR-NILMRC-194/2020\|EPI_ISL_4 | ···················· | ···················· | ···················· | ···················· | ···················· | ···················· |
| E | 8843 | hCoV-19/USA/WI-UW-595/2020\|EPI_ISL_484970\|2020-06- | ···················· | ···················· | ···················· | ···················· | ···················· | ···················· |
| E | 8844 | hCoV-19/USA/WI-UW-596/2020\|EPI_ISL_484971\|2020-06- | ···················· | ···················· | ···················· | ···················· | ···················· | ···················· |
| E | 8845 | hCoV-19/USA/NY-UW-597/2020\|EPI_ISL_484972\|2020-06- | ···················· | ···················· | ···················· | ···················· | ···················· | ···················· |
| E | 8846 | hCoV-19/USA/WI-UW-598/2020\|EPI_ISL_484973\|2020-06- | ···················· | ···················· | ···················· | ···················· | ···················· | ···················· |
| E | 8847 | hCoV-19/USA/WI-UW-599/2020\|EPI_ISL_484974\|2020-06- | ···················· | ···················· | ···················· | ···················· | ···················· | ···················· |
| E | 8848 | hCoV-19/USA/IL-UW-600/2020\|EPI_ISL_484975\|2020-06- | ···················· | ···················· | ···················· | ···················· | ···················· | ···················· |
| E | 8849 | hCoV-19/USA/WI-UW-601/2020\|EPI_ISL_484976\|2020-06- | ···················· | ···················· | ···················· | ···················· | ···················· | ···················· |
| E | 8850 | hCoV-19/USA/WI-UW-603/2020\|EPI_ISL_484978\|2020-06- | ···················· | ···················· | ···················· | ···················· | ···················· | ···················· |
| E | 8851 | hCoV-19/USA/WI-UW-604/2020\|EPI_ISL_484979\|2020-06- | ···················· | ···················· | ···················· | ···················· | ···················· | ···················· |
| E | 8852 | hCoV-19/USA/CA-UW-606/2020\|EPI_ISL_484981\|2020-06- | ···················· | ···················· | ···················· | ···················· | ···················· | ···················· |
| E | 8856 | hCoV-19/USA/WI-UW-610/2020\|EPI_ISL_484985\|2020-06- | ···················· | ···················· | ···················· | ···················· | ···················· | ···················· |
| E | 8857 | hCoV-19/USA/WI-UW-611/2020\|EPI_ISL_484986\|2020-06- | ···················· | ···················· | ···················· | ···················· | ···················· | ···················· |
| E | 8858 | hCoV-19/USA/WI-UW-612/2020\|EPI_ISL_484987\|2020-06- | ···················· | ···················· | ···················· | ···················· | ···················· | ···················· |

| | | | | | | | | |
|---|---|---|---|---|---|---|---|---|
| E | 8859 | hCoV-19/USA/WI-UW-613/2020\|EPI_ISL_484988\|2020-06- | ···················· | ···················· | ···················· | ···················· | ···················· | ···················· |
| E | 8860 | hCoV-19/USA/WI-UW-614/2020\|EPI_ISL_484989\|2020-06- | ···················· | ···················· | ···················· | ···················· | ···················· | ···················· |
| E | 8861 | hCoV-19/USA/WI-UW-615/2020\|EPI_ISL_484990\|2020-06- | ···················· | ···················· | ···················· | ···················· | ···················· | ···················· |
| E | 8862 | hCoV-19/USA/WI-UW-616/2020\|EPI_ISL_484991\|2020-06- | ···················· | ···················· | ···················· | ···················· | ···················· | ···················· |
| E | 8863 | hCoV-19/USA/WI-UW-617/2020\|EPI_ISL_484992\|2020-06- | ···················· | ···················· | ···················· | ···················· | ···················· | ···················· |
| E | 8864 | hCoV-19/USA/WI-UW-618/2020\|EPI_ISL_484993\|2020-06- | ···················· | ···················· | ···················· | ···················· | ···················· | ···················· |
| E | 8865 | hCoV-19/USA/WI-UW-619/2020\|EPI_ISL_484994\|2020-06- | ···················· | ···················· | ···················· | ···················· | ···················· | ···················· |
| E | 8866 | hCoV-19/USA/WI-UW-620/2020\|EPI_ISL_484995\|2020-06- | ···················· | ···················· | ···················· | ···················· | ···················· | ···················· |
| E | 8867 | hCoV-19/USA/WI-UW-622/2020\|EPI_ISL_484997\|2020-06- | ···················· | ···················· | ···················· | ···················· | ···················· | ···················· |
| E | 8868 | hCoV-19/USA/WI-UW-623/2020\|EPI_ISL_484998\|2020-06- | ···················· | ···················· | ···················· | ···················· | ···················· | ···················· |
| E | 8869 | hCoV-19/USA/WI-UW-624/2020\|EPI_ISL_484999\|2020-06- | ···················· | ···················· | ···················· | ···················· | ···················· | ···················· |
| E | 8870 | hCoV-19/Latvia/065/2020\|EPI_ISL_493000\|2020-07-09 | ···················· | ···················· | ···················· | ···················· | ···················· | ···················· |
| E | 8871 | hCoV-19/USA/WY-WYPHL-20007938/2020\|EPI_ISL_493001\| | ···················· | ···················· | ···················· | ···················· | ···················· | ···················· |
| E | 8872 | hCoV-19/England/20284212204/2020\|EPI_ISL_493002\|20 | ···················· | ···················· | ···················· | ···················· | ···················· | ···················· |
| E | 8873 | hCoV-19/England/20284212304/2020\|EPI_ISL_493003\|20 | ···················· | ···················· | ···················· | ···················· | ···················· | ···················· |
| E | 8874 | hCoV-19/USA/UT-03265/2020\|EPI_ISL_493006\|2020-05-0 | ···················· | ···················· | ···················· | ···················· | ···················· | ···················· |
| E | 8875 | hCoV-19/USA/UT-03267/2020\|EPI_ISL_493007\|2020-05-0 | ···················· | ···················· | ···················· | ···················· | ···················· | ···················· |
| E | 8876 | hCoV-19/USA/UT-03270/2020\|EPI_ISL_493009\|2020-05-0 | ···················· | ···················· | ···················· | ···················· | ···················· | ···················· |
| E | 8877 | hCoV-19/USA/UT-03272/2020\|EPI_ISL_493010\|2020-05-0 | ···················· | ···················· | ···················· | ···················· | ···················· | ···················· |
| E | 8878 | hCoV-19/USA/UT-03826/2020\|EPI_ISL_493022\|2020-05-0 | ···················· | ···················· | ···················· | ···················· | ···················· | ···················· |
| E | 8879 | hCoV-19/USA/UT-03827/2020\|EPI_ISL_493023\|2020-05-0 | ···················· | ···················· | ···················· | ···················· | ···················· | ···················· |
| E | 8880 | hCoV-19/USA/UT-04131/2020\|EPI_ISL_493030\|2020-05-1 | ···················· | ···················· | ···················· | ···················· | ···················· | ···················· |
| E | 8881 | hCoV-19/USA/UT-04137/2020\|EPI_ISL_493036\|2020-05-1 | ···················· | ···················· | ···················· | ···················· | ···················· | ···················· |
| E | 8882 | hCoV-19/USA/UT-04138/2020\|EPI_ISL_493037\|2020-05-1 | ···················· | ···················· | ···················· | ···················· | ···················· | ···················· |
| E | 8883 | hCoV-19/USA/UT-04143/2020\|EPI_ISL_493041\|2020-05-1 | ···················· | ···················· | ···················· | ···················· | ···················· | ···················· |
| E | 8884 | hCoV-19/USA/UT-04149/2020\|EPI_ISL_493042\|2020-05-1 | ···················· | ···················· | ···················· | ···················· | ···················· | ···················· |
| E | 8885 | hCoV-19/Wales/PHWC-1688FE/2020\|EPI_ISL_494303\|2020 | ···················· | ···················· | ···················· | ···················· | ···················· | ···················· |
| E | 8886 | hCoV-19/Wales/PHWC-168919/2020\|EPI_ISL_494305\|2020 | ···················· | ···················· | ···················· | ···················· | ···················· | ···················· |
| E | 8887 | hCoV-19/Wales/PHWC-168946/2020\|EPI_ISL_494308\|2020 | ···················· | ···················· | ···················· | ···················· | ···················· | ···················· |
| E | 8888 | hCoV-19/Wales/PHWC-168964/2020\|EPI_ISL_494309\|2020 | ···················· | ···················· | ···················· | ···················· | ···················· | ···················· |
| E | 8889 | hCoV-19/Wales/PHWC-1689A0/2020\|EPI_ISL_494312\|2020 | ···················· | ···················· | ···················· | ···················· | ···················· | ···················· |
| E | 8890 | hCoV-19/Wales/PHWC-168A43/2020\|EPI_ISL_494319\|2020 | ···················· | ···················· | ···················· | ···················· | ···················· | ···················· |
| E | 8891 | hCoV-19/Wales/PHWC-168A61/2020\|EPI_ISL_494321\|2020 | ···················· | ····A··············· | ···················· | ···················· | ···················· | ···················· |
| E | 8892 | hCoV-19/Wales/PHWC-168ACB/2020\|EPI_ISL_494326\|2020 | ···················· | ···················· | ···················· | ···················· | ···················· | ···················· |
| E | 8893 | hCoV-19/Wales/PHWC-168B22/2020\|EPI_ISL_494332\|2020 | ···················· | ···················· | ···················· | ···················· | ···················· | ···················· |
| E | 8894 | hCoV-19/Wales/PHWC-168B6E/2020\|EPI_ISL_494336\|2020 | ···················· | ···················· | ···················· | ···················· | ···················· | ···················· |
| E | 8895 | hCoV-19/Wales/PHWC-168B8C/2020\|EPI_ISL_494338\|2020 | ···················· | ···················· | ···················· | ···················· | ···················· | ···················· |
| E | 8896 | hCoV-19/Wales/PHWC-168BB9/2020\|EPI_ISL_494341\|2020 | ···················· | ···················· | ···················· | ···················· | ···················· | ···················· |
| E | 8897 | hCoV-19/Wales/PHWC-168D4A/2020\|EPI_ISL_494359\|2020 | ···················· | ···················· | ···················· | ···················· | ···················· | ···················· |
| E | 8898 | hCoV-19/Wales/PHWC-168D59/2020\|EPI_ISL_494360\|2020 | ···················· | ···················· | ···················· | ···················· | ···················· | ···················· |
| E | 8899 | hCoV-19/Wales/PHWC-168D68/2020\|EPI_ISL_494361\|2020 | ···················· | ···················· | ···················· | ···················· | ···················· | ···················· |
| E | 8900 | hCoV-19/Wales/PHWC-168D95/2020\|EPI_ISL_494364\|2020 | ···················· | ···················· | ···················· | ···················· | ···················· | ···················· |
| E | 8901 | hCoV-19/Wales/PHWC-168DE0/2020\|EPI_ISL_494368\|2020 | ···················· | ···················· | ···················· | ···················· | ···················· | ···················· |
| E | 8902 | hCoV-19/Wales/PHWC-168E29/2020\|EPI_ISL_494370\|2020 | ···················· | ···················· | ···················· | ···················· | ···················· | ···················· |
| E | 8903 | hCoV-19/USA/CA-ALSR-1119/2020\|EPI_ISL_494372\|2020- | ···················· | ···················· | ···················· | ···················· | ···················· | ···················· |
| E | 8904 | hCoV-19/USA/CA-ALSR-1120/2020\|EPI_ISL_494373\|2020- | ···················· | ···················· | ···················· | ···················· | ···················· | ···················· |
| E | 8905 | hCoV-19/USA/CA-ALSR-1121/2020\|EPI_ISL_494374\|2020- | ···················· | ···················· | ···················· | ···················· | ···················· | ···················· |
| E | 8906 | hCoV-19/USA/MN-MDH-1321/2020\|EPI_ISL_495620\|2020-0 | ···················· | ···················· | ···················· | ···················· | ···················· | ···················· |
| E | 8907 | hCoV-19/USA/MN-MDH-1322/2020\|EPI_ISL_495621\|2020-0 | ···················· | ···················· | ···················· | ···················· | ···················· | ···················· |
| E | 8908 | hCoV-19/USA/MN-MDH-1323/2020\|EPI_ISL_495622\|2020-0 | ···················· | ···················· | ···················· | ···················· | ···················· | ···················· |
| E | 8909 | hCoV-19/USA/MN-MDH-1324/2020\|EPI_ISL_495623\|2020-0 | ···················· | ···················· | ···················· | ···················· | ···················· | ···················· |
| E | 8910 | hCoV-19/USA/MN-MDH-1325/2020\|EPI_ISL_495624\|2020-0 | ···················· | ···················· | ···················· | ···················· | ···················· | ···················· |
| E | 8911 | hCoV-19/USA/MN-MDH-1326/2020\|EPI_ISL_495625\|2020-0 | ···················· | ···················· | ···················· | ···················· | ···················· | ···················· |
| E | 8912 | hCoV-19/USA/MN-MDH-1327/2020\|EPI_ISL_495626\|2020-0 | ···················· | ···················· | ···················· | ···················· | ···················· | ···················· |
| E | 8913 | hCoV-19/USA/MN-MDH-1328/2020\|EPI_ISL_495627\|2020-0 | ···················· | ···················· | ···················· | ···················· | ···················· | ···················· |
| E | 8914 | hCoV-19/USA/MN-MDH-1329/2020\|EPI_ISL_495628\|2020-0 | ···················· | ···················· | ···················· | ···················· | ···················· | ···················· |
| E | 8915 | hCoV-19/DRC/6724/2020\|EPI_ISL_495634\|2020-05-11 | ···················· | ···················· | ···················· | ···················· | ···················· | ···················· |
| E | 8916 | hCoV-19/DRC/6784/2020\|EPI_ISL_495636\|2020-05-11 | ···················· | ···················· | ···················· | ···················· | ···················· | ···················· |
| E | 8917 | hCoV-19/DRC/6786/2020\|EPI_ISL_495637\|2020-05-11 | ···················· | ···················· | ···················· | ···················· | ···················· | ···················· |
| E | 8918 | hCoV-19/DRC/7920/2020\|EPI_ISL_495641\|2020-05-15 | ···················· | ···················· | ···················· | ···················· | ···················· | ···················· |
| E | 8919 | hCoV-19/USA/WA-S1467/2020\|EPI_ISL_495642\|2020-05-0 | ···················· | ···················· | ···················· | ···················· | ···················· | ···················· |
| E | 8920 | hCoV-19/USA/WA-S1468/2020\|EPI_ISL_495643\|2020-05-1 | ···················· | ···················· | ···················· | ···················· | ···················· | ···················· |
| E | 8921 | hCoV-19/USA/WA-S1469/2020\|EPI_ISL_495644\|2020-05-0 | ···················· | ···················· | ···················· | ···················· | ···················· | ···················· |
| E | 8922 | hCoV-19/USA/WA-S1471/2020\|EPI_ISL_495646\|2020-05-0 | ···················· | ···················· | ···················· | ···················· | ···················· | ···················· |
| E | 8923 | hCoV-19/USA/WA-S1472/2020\|EPI_ISL_495647\|2020-05-0 | ···················· | ···················· | ···················· | ···················· | ···················· | ···················· |
| E | 8924 | hCoV-19/USA/WA-S1473/2020\|EPI_ISL_495648\|2020-05-1 | ···················· | ···················· | ···················· | ···················· | ···················· | ···················· |
| E | 8925 | hCoV-19/USA/WA-S1474/2020\|EPI_ISL_495649\|2020-05-1 | ···················· | ···················· | ···················· | ···················· | ···················· | ···················· |
| E | 8926 | hCoV-19/USA/WA-S1476/2020\|EPI_ISL_495651\|2020-05-1 | ···················· | ···················· | ···················· | ···················· | ···················· | ···················· |
| E | 8927 | hCoV-19/USA/WA-S1477/2020\|EPI_ISL_495652\|2020-05-1 | ···················· | ···················· | ···················· | ···················· | ···················· | ···················· |
| E | 8928 | hCoV-19/USA/WA-S1478/2020\|EPI_ISL_495653\|2020-05-1 | ···················· | ···················· | ···················· | ···················· | ···················· | ···················· |
| E | 8929 | hCoV-19/USA/WA-S1479/2020\|EPI_ISL_495654\|2020-05-0 | ···················· | ···················· | ···················· | ···················· | ···················· | ···················· |
| E | 8930 | hCoV-19/USA/WA-S1480/2020\|EPI_ISL_495655\|2020-05-0 | ···················· | ···················· | ···················· | ···················· | ···················· | ···················· |
| E | 8931 | hCoV-19/USA/WA-S1482/2020\|EPI_ISL_495657\|2020-03-2 | ···················· | ···················· | ···················· | ···················· | ···················· | ···················· |
| E | 8932 | hCoV-19/USA/WA-S1483/2020\|EPI_ISL_495658\|2020-03-2 | ···················· | ···················· | ···················· | ···················· | ···················· | ···················· |
| E | 8933 | hCoV-19/USA/WA-S1484/2020\|EPI_ISL_495659\|2020-05-0 | ···················· | ···················· | ···················· | ···················· | ···················· | ···················· |
| E | 8934 | hCoV-19/USA/WA-S1485/2020\|EPI_ISL_495660\|2020-05-0 | ···················· | ···················· | ···················· | ···················· | ···················· | ···················· |
| E | 8935 | hCoV-19/USA/WA-S1486/2020\|EPI_ISL_495661\|2020-03-2 | ···················· | ···················· | ···················· | ···················· | ···················· | ···················· |
| E | 8936 | hCoV-19/USA/WA-S1487/2020\|EPI_ISL_495662\|2020-05-0 | ···················· | ···················· | ···················· | ···················· | ···················· | ···················· |
| E | 8937 | hCoV-19/USA/WA-S1488/2020\|EPI_ISL_495663\|2020-03-0 | ···················· | ···················· | ···················· | ···················· | ···················· | ···················· |
| E | 8938 | hCoV-19/USA/WA-S1489/2020\|EPI_ISL_495664\|2020-05-0 | ···················· | ···················· | ···················· | ···················· | ···················· | ···················· |

| | | | | | | | |
|---|---|---|---|---|---|---|---|
| E | 8939 hCoV-19/USA/WA-S1490/2020|EPI_ISL_495665|2020-05-0 | .................... | .................... | .................... | .................... | .................... | .................... |
| E | 8940 hCoV-19/USA/WA-S1216/2020|EPI_ISL_495666|2020-05-0 | .................... | .................... | .................... | .................... | .................... | .................... |
| E | 8941 hCoV-19/USA/WA-S1217/2020|EPI_ISL_495667|2020-05-0 | .................... | .................... | .................... | .................... | .................... | .................... |
| E | 8942 hCoV-19/USA/WA-S1218/2020|EPI_ISL_495668|2020-05-0 | .................... | .................... | .................... | .................... | .................... | .................... |
| E | 8943 hCoV-19/USA/WA-S1219/2020|EPI_ISL_495669|2020-05-0 | .................... | .................... | .................... | .................... | .................... | .................... |
| E | 8944 hCoV-19/USA/WA-S1220/2020|EPI_ISL_495670|2020-05-0 | .................... | .................... | .................... | .................... | .................... | .................... |
| E | 8945 hCoV-19/USA/WA-S1221/2020|EPI_ISL_495671|2020-05-0 | .................... | .................... | .................... | .................... | .................... | .................... |
| E | 8946 hCoV-19/USA/WA-S1222/2020|EPI_ISL_495672|2020-05-0 | .................... | .................... | .................... | .................... | .................... | .................... |
| E | 8947 hCoV-19/USA/WA-S1234/2020|EPI_ISL_495684|2020-05-1 | .................... | .................... | .................... | .................... | .................... | .................... |
| E | 8948 hCoV-19/USA/WA-S1235/2020|EPI_ISL_495685|2020-05-0 | .................... | .................... | .................... | .................... | .................... | .................... |
| E | 8949 hCoV-19/USA/WA-S1236/2020|EPI_ISL_495686|2020-05-0 | .................... | .................... | .................... | .................... | .................... | .................... |
| E | 8950 hCoV-19/USA/WA-S1237/2020|EPI_ISL_495687|2020-05-1 | .................... | .................... | .................... | .................... | .................... | .................... |
| E | 8951 hCoV-19/USA/WA-S1238/2020|EPI_ISL_495688|2020-05-1 | .................... | .................... | .................... | .................... | .................... | .................... |
| E | 8952 hCoV-19/USA/WA-S1239/2020|EPI_ISL_495689|2020-05-1 | .................... | .................... | .................... | .................... | .................... | .................... |
| E | 8953 hCoV-19/USA/WA-S1240/2020|EPI_ISL_495690|2020-05-1 | .................... | .................... | .................... | .................... | .................... | .................... |
| E | 8954 hCoV-19/USA/WA-S1242/2020|EPI_ISL_495692|2020-05-1 | .................... | .................... | .................... | .................... | .................... | .................... |
| E | 8955 hCoV-19/USA/WA-S1243/2020|EPI_ISL_495693|2020-05-1 | .................... | .................... | .................... | .................... | .................... | .................... |
| E | 8956 hCoV-19/USA/WA-S1244/2020|EPI_ISL_495694|2020-05-1 | .................... | .................... | .................... | .................... | .................... | .................... |
| E | 8957 hCoV-19/USA/WA-S1245/2020|EPI_ISL_495695|2020-05-1 | .................... | .................... | .................... | .................... | .................... | .................... |
| E | 8958 hCoV-19/USA/WA-S1246/2020|EPI_ISL_495696|2020-05-1 | .................... | .................... | .................... | .................... | .................... | .................... |
| E | 8959 hCoV-19/USA/WA-S1247/2020|EPI_ISL_495697|2020-05-1 | .................... | .................... | .................... | .................... | .................... | .................... |
| E | 8960 hCoV-19/USA/WA-S1248/2020|EPI_ISL_495698|2020-05-1 | .................... | .................... | .................... | .................... | .................... | .................... |
| E | 8965 hCoV-19/USA/OR-PROV-013/2020|EPI_ISL_482300|2020-0 | .................... | .................... | .................... | .................... | .................... | .................... |
| E | 8966 hCoV-19/USA/OR-PROV-014/2020|EPI_ISL_482301|2020-0 | .................... | .................... | .................... | .................... | .................... | .................... |
| E | 8967 hCoV-19/USA/OR-PROV-015/2020|EPI_ISL_482302|2020-0 | .................... | .................... | .................... | .................... | .................... | .................... |
| E | 8968 hCoV-19/USA/OR-PROV-016/2020|EPI_ISL_482303|2020-0 | .................... | .................... | .................... | .................... | .................... | .................... |
| E | 8969 hCoV-19/USA/OR-PROV-017/2020|EPI_ISL_482304|2020-0 | .................... | .................... | .................... | .................... | .................... | .................... |
| E | 8970 hCoV-19/USA/OR-PROV-018/2020|EPI_ISL_482305|2020-0 | .................... | .................... | .................... | .................... | .................... | .................... |
| E | 8971 hCoV-19/USA/OR-PROV-020/2020|EPI_ISL_482306|2020-0 | .................... | .................... | .................... | .................... | .................... | .................... |
| E | 8972 hCoV-19/USA/OR-PROV-023/2020|EPI_ISL_482307|2020-0 | .................... | .................... | .................... | .................... | .................... | .................... |
| E | 8973 hCoV-19/USA/OR-PROV-024/2020|EPI_ISL_482308|2020-0 | .................... | .................... | .................... | .................... | .................... | .................... |
| E | 8974 hCoV-19/USA/OR-PROV-025/2020|EPI_ISL_482309|2020-0 | .................... | .................... | .................... | .................... | .................... | .................... |
| E | 8975 hCoV-19/USA/OR-PROV-066/2020|EPI_ISL_482345|2020-0 | .................... | .................... | .................... | .................... | .................... | .................... |
| E | 8976 hCoV-19/USA/OR-PROV-068/2020|EPI_ISL_482346|2020-0 | .................... | .................... | .................... | .................... | .................... | .................... |
| E | 8977 hCoV-19/USA/OR-PROV-074/2020|EPI_ISL_482349|2020-0 | .................... | .................... | .................... | .................... | .................... | .................... |
| E | 8978 hCoV-19/Singapore/577/2020|EPI_ISL_483600|2020-04- | .................... | .................... | .................... | .................... | .................... | .................... |
| E | 8979 hCoV-19/Singapore/578/2020|EPI_ISL_483601|2020-04- | .................... | .................... | .................... | .................... | .................... | .................... |
| E | 8980 hCoV-19/Singapore/579/2020|EPI_ISL_483602|2020-04- | .................... | .................... | .................... | .................... | .................... | .................... |
| E | 8981 hCoV-19/Singapore/580/2020|EPI_ISL_483603|2020-05- | .................... | .................... | .................... | .................... | .................... | .................... |
| E | 8982 hCoV-19/Singapore/581/2020|EPI_ISL_483604|2020-04- | .................... | .................... | .................... | .................... | .................... | .................... |
| E | 8983 hCoV-19/Singapore/582/2020|EPI_ISL_483605|2020-04- | .................... | .................... | .................... | .................... | .................... | .................... |
| E | 8984 hCoV-19/Singapore/583/2020|EPI_ISL_483606|2020-05- | .................... | .................... | .................... | .................... | .................... | .................... |
| E | 8985 hCoV-19/Singapore/584/2020|EPI_ISL_483607|2020-05- | .................... | .................... | .................... | .................... | .................... | .................... |
| E | 8986 hCoV-19/Singapore/585/2020|EPI_ISL_483608|2020-06- | .................... | .................... | .................... | .................... | .................... | .................... |
| E | 8987 hCoV-19/Singapore/586/2020|EPI_ISL_483609|2020-04- | .................... | .................... | .................... | .................... | .................... | .................... |
| E | 8988 hCoV-19/Singapore/587/2020|EPI_ISL_483610|2020-03- | .................... | .................... | .................... | .................... | .................... | .................... |
| E | 8989 hCoV-19/Singapore/588/2020|EPI_ISL_483611|2020-06- | .................... | .................... | .................... | .................... | .................... | .................... |
| E | 8990 hCoV-19/Singapore/589/2020|EPI_ISL_483612|2020-04- | .................... | .................... | .................... | .................... | .................... | .................... |
| E | 8991 hCoV-19/Singapore/590/2020|EPI_ISL_483613|2020-03- | .................... | .................... | .................... | .................... | .................... | .................... |
| E | 8992 hCoV-19/Singapore/591/2020|EPI_ISL_483614|2020-06- | .................... | .................... | .................... | .................... | .................... | .................... |
| E | 8993 hCoV-19/Singapore/592/2020|EPI_ISL_483615|2020-06- | .................... | .................... | .................... | .................... | .................... | .................... |
| E | 8994 hCoV-19/Singapore/593/2020|EPI_ISL_483616|2020-05- | .................... | .................... | .................... | .................... | .................... | .................... |
| E | 8995 hCoV-19/Singapore/594/2020|EPI_ISL_483617|2020-03- | .................... | .................... | .................... | .................... | .................... | .................... |
| E | 8996 hCoV-19/Singapore/595/2020|EPI_ISL_483618|2020-06- | .................... | .................... | .................... | .................... | .................... | .................... |
| E | 8997 hCoV-19/Singapore/596/2020|EPI_ISL_483619|2020-06- | .................... | .................... | .................... | .................... | .................... | .................... |
| E | 8998 hCoV-19/Singapore/597/2020|EPI_ISL_483620|2020-06- | .................... | .................... | .................... | .................... | .................... | .................... |
| E | 8999 hCoV-19/Singapore/598/2020|EPI_ISL_483621|2020-03- | .................... | .................... | .................... | .................... | .................... | .................... |
| E | 9000 hCoV-19/Bangladesh/BCSIR-NILMRC-175/2020|EPI_ISL_4 | .................... | .................... | .................... | .................... | .................... | .................... |
| E | 9001 hCoV-19/Bangladesh/BCSIR-NILMRC-176/2020|EPI_ISL_4 | .................... | .................... | .................... | .................... | .................... | .................... |
| E | 9002 hCoV-19/Bangladesh/BCSIR-NILMRC-177/2020|EPI_ISL_4 | .................... | .................... | .................... | .................... | .................... | .................... |
| E | 9003 hCoV-19/Bangladesh/BCSIR-NILMRC-178/2020|EPI_ISL_4 | .................... | .................... | .................... | .................... | .................... | .................... |
| E | 9004 hCoV-19/Bangladesh/BCSIR-NILMRC-174/2020|EPI_ISL_4 | .................... | .................... | .................... | .................... | .................... | .................... |
| E | 9005 hCoV-19/Bangladesh/BCSIR-NILMRC-179/2020|EPI_ISL_4 | .................... | .................... | .................... | .................... | .................... | .................... |
| E | 9006 hCoV-19/Bangladesh/BCSIR-NILMRC-181/2020|EPI_ISL_4 | .................... | .................... | .................... | .................... | .................... | .................... |
| E | 9007 hCoV-19/Bangladesh/BCSIR-NILMRC-182/2020|EPI_ISL_4 | .................... | .................... | .................... | .................... | .................... | .................... |
| E | 9008 hCoV-19/Bangladesh/BCSIR-NILMRC-184/2020|EPI_ISL_4 | .................... | .................... | .................... | .................... | .................... | .................... |
| E | 9009 hCoV-19/Bangladesh/BCSIR-NILMRC-187/2020|EPI_ISL_4 | .................... | .................... | .................... | .................... | .................... | .................... |
| E | 9010 hCoV-19/Bangladesh/BCSIR-NILMRC-188/2020|EPI_ISL_4 | .................... | .................... | .................... | .................... | .................... | .................... |
| E | 9011 hCoV-19/Bangladesh/BCSIR-NILMRC-192/2020|EPI_ISL_4 | .................... | .................... | .................... | .................... | .................... | .................... |
| E | 9012 hCoV-19/Bangladesh/BCSIR-NILMRC-193/2020|EPI_ISL_4 | .................... | .................... | .................... | .................... | .................... | .................... |
| E | 9013 hCoV-19/Bangladesh/BCSIR-NILMRC-196/2020|EPI_ISL_4 | .................... | .................... | .................... | .................... | .................... | .................... |
| E | 9014 hCoV-19/Hungary/SRC_isolate_2/2020|EPI_ISL_483637| | .................... | .................... | .................... | .................... | .................... | .................... |
| E | 9015 hCoV-19/Bahrein/BAH-25/2020|EPI_ISL_483639|2020-04 | .................... | .................... | .................... | .................... | .................... | .................... |
| E | 9016 hCoV-19/Switzerland/ZH-ETHZ-768plus/2020|EPI_ISL_4 | .................... | .................... | .................... | .................... | .................... | .................... |
| E | 9017 hCoV-19/USA/IL-UW-525/2020|EPI_ISL_484900|2020-06- | .................... | .................... | .................... | .................... | .................... | .................... |
| E | 9018 hCoV-19/USA/WI-UW-526/2020|EPI_ISL_484901|2020-06- | .................... | .................... | .................... | .................... | .................... | .................... |
| E | 9019 hCoV-19/USA/WI-UW-527/2020|EPI_ISL_484902|2020-06- | .................... | .................... | .................... | .................... | .................... | .................... |
| E | 9020 hCoV-19/USA/WI-UW-528/2020|EPI_ISL_484903|2020-06- | .................... | .................... | .................... | .................... | .................... | .................... |
| E | 9021 hCoV-19/USA/WI-UW-529/2020|EPI_ISL_484904|2020-06- | .................... | .................... | .................... | .................... | .................... | .................... |
| E | 9022 hCoV-19/USA/WI-UW-530/2020|EPI_ISL_484905|2020-06- | .................... | .................... | .................... | .................... | .................... | .................... |

| | | | | | | | | | |
|---|---|---|---|---|---|---|---|---|---|
| E | 9023 | hCoV-19/USA/IL-UW-531/2020\|EPI_ISL_484906\|2020-06- | ···················· | ···················· | ···················· | ···················· | ···················· | ···················· | |
| E | 9024 | hCoV-19/USA/IL-UW-532/2020\|EPI_ISL_484907\|2020-06- | ···················· | ···················· | ···················· | ···················· | ···················· | ···················· | |
| E | 9025 | hCoV-19/USA/WI-UW-533/2020\|EPI_ISL_484908\|2020-06- | ···················· | ···················· | ···················· | ···················· | ···················· | ···················· | |
| E | 9026 | hCoV-19/USA/WI-UW-534/2020\|EPI_ISL_484909\|2020-06- | ···················· | ···················· | ···················· | ···················· | ···················· | ···················· | |
| E | 9027 | hCoV-19/USA/WI-UW-535/2020\|EPI_ISL_484910\|2020-06- | ···················· | ···················· | ···················· | ···················· | ···················· | ···················· | |
| E | 9028 | hCoV-19/USA/WI-UW-536/2020\|EPI_ISL_484911\|2020-06- | ···················· | ···················· | ···················· | ···················· | ···················· | ···················· | |
| E | 9032 | hCoV-19/USA/WI-UW-541/2020\|EPI_ISL_484916\|2020-06- | ···················· | ···················· | ···················· | ···················· | ···················· | ···················· | |
| E | 9033 | hCoV-19/USA/IL-UW-542/2020\|EPI_ISL_484917\|2020-06- | ···················· | ···················· | ···················· | ···················· | ···················· | ···················· | |
| E | 9035 | hCoV-19/USA/WI-UW-544/2020\|EPI_ISL_484919\|2020-06- | ···················· | ···················· | ···················· | ···················· | ···················· | ···················· | |
| E | 9038 | hCoV-19/USA/PA-UW-547/2020\|EPI_ISL_484922\|2020-06- | ···················· | ···················· | ···················· | ···················· | ···················· | ···················· | |
| E | 9041 | hCoV-19/USA/WI-UW-551/2020\|EPI_ISL_484926\|2020-06- | ···················· | ···················· | ···················· | ···················· | ···················· | ···················· | |
| E | 9042 | hCoV-19/USA/WI-UW-552/2020\|EPI_ISL_484927\|2020-06- | ···················· | ···················· | ···················· | ···················· | ···················· | ···················· | |
| E | 9044 | hCoV-19/USA/WI-UW-556/2020\|EPI_ISL_484931\|2020-06- | ···················· | ···················· | ···················· | ···················· | ···················· | ···················· | |
| E | 9045 | hCoV-19/USA/NJ-UW-557/2020\|EPI_ISL_484932\|2020-06- | ···················· | ···················· | ···················· | ···················· | ···················· | ···················· | |
| E | 9048 | hCoV-19/USA/WI-UW-560/2020\|EPI_ISL_484935\|2020-06- | ···················· | ···················· | ···················· | ···················· | ···················· | ···················· | |
| E | 9049 | hCoV-19/USA/WI-UW-561/2020\|EPI_ISL_484936\|2020-06- | ···················· | ···················· | ···················· | ···················· | ···················· | ···················· | |
| E | 9051 | hCoV-19/USA/WI-UW-563/2020\|EPI_ISL_484938\|2020-06- | ···················· | ···················· | ···················· | ···················· | ···················· | ···················· | |
| E | 9052 | hCoV-19/USA/WI-UW-564/2020\|EPI_ISL_484939\|2020-06- | ···················· | ···················· | ···················· | ···················· | ···················· | ···················· | |
| E | 9053 | hCoV-19/USA/WI-UW-565/2020\|EPI_ISL_484940\|2020-06- | ···················· | ···················· | ···················· | ···················· | ···················· | ···················· | |
| E | 9054 | hCoV-19/USA/WI-UW-566/2020\|EPI_ISL_484941\|2020-06- | ···················· | ···················· | ···················· | ···················· | ···················· | ···················· | |
| E | 9055 | hCoV-19/USA/WI-UW-567/2020\|EPI_ISL_484942\|2020-06- | ···················· | ···················· | ···················· | ···················· | ···················· | ···················· | |
| E | 9056 | hCoV-19/USA/WI-UW-569/2020\|EPI_ISL_484944\|2020-06- | ···················· | ···················· | ···················· | ···················· | ···················· | ···················· | |
| E | 9057 | hCoV-19/USA/WI-UW-570/2020\|EPI_ISL_484945\|2020-06- | ···················· | ···················· | ···················· | ···················· | ···················· | ···················· | |
| E | 9058 | hCoV-19/USA/IL-UW-571/2020\|EPI_ISL_484946\|2020-06- | ···················· | ···················· | ···················· | ···················· | ···················· | ···················· | |
| E | 9059 | hCoV-19/USA/WI-UW-572/2020\|EPI_ISL_484947\|2020-06- | ···················· | ···················· | ···················· | ···················· | ···················· | ···················· | |
| E | 9060 | hCoV-19/USA/WI-UW-573/2020\|EPI_ISL_484948\|2020-06- | ···················· | ···················· | ···················· | ···················· | ···················· | ···················· | |
| E | 9061 | hCoV-19/USA/WI-UW-574/2020\|EPI_ISL_484949\|2020-06- | ···················· | ···················· | ···················· | ···················· | ···················· | ···················· | |
| E | 9062 | hCoV-19/USA/WI-UW-575/2020\|EPI_ISL_484950\|2020-06- | ···················· | ···················· | ···················· | ···················· | ···················· | ···················· | |
| E | 9063 | hCoV-19/USA/WI-UW-576/2020\|EPI_ISL_484951\|2020-06- | ···················· | ···················· | ···················· | ···················· | ···················· | ···················· | |
| E | 9064 | hCoV-19/USA/WI-UW-577/2020\|EPI_ISL_484952\|2020-06- | ···················· | ···················· | ···················· | ···················· | ···················· | ···················· | |
| E | 9065 | hCoV-19/USA/WI-UW-578/2020\|EPI_ISL_484953\|2020-06- | ···················· | ···················· | ···················· | ···················· | ···················· | ···················· | |
| E | 9066 | hCoV-19/USA/WI-UW-579/2020\|EPI_ISL_484954\|2020-06- | ···················· | ···················· | ···················· | ···················· | ···················· | ···················· | |
| E | 9067 | hCoV-19/USA/WI-UW-580/2020\|EPI_ISL_484955\|2020-06- | ···················· | ···················· | ···················· | ···················· | ···················· | ···················· | |
| E | 9068 | hCoV-19/USA/WI-UW-581/2020\|EPI_ISL_484956\|2020-06- | ···················· | ···················· | ···················· | ···················· | ···················· | ···················· | |
| E | 9069 | hCoV-19/USA/WI-UW-582/2020\|EPI_ISL_484957\|2020-06- | ···················· | ···················· | ···················· | ···················· | ···················· | ···················· | |
| E | 9070 | hCoV-19/USA/WI-UW-583/2020\|EPI_ISL_484958\|2020-06- | ···················· | ···················· | ···················· | ···················· | ···················· | ···················· | |
| E | 9071 | hCoV-19/USA/NY-UW-584/2020\|EPI_ISL_484959\|2020-06- | ···················· | ···················· | ···················· | ···················· | ···················· | ···················· | |
| E | 9072 | hCoV-19/USA/WI-UW-585/2020\|EPI_ISL_484960\|2020-06- | ···················· | ···················· | ···················· | ···················· | ···················· | ···················· | |
| E | 9073 | hCoV-19/USA/WI-UW-586/2020\|EPI_ISL_484961\|2020-06- | ···················· | ···················· | ···················· | ···················· | ···················· | ···················· | |
| E | 9074 | hCoV-19/USA/WI-UW-587/2020\|EPI_ISL_484962\|2020-06- | ···················· | ···················· | ···················· | ···················· | ···················· | ···················· | |
| E | 9075 | hCoV-19/USA/WI-UW-588/2020\|EPI_ISL_484963\|2020-06- | ···················· | ···················· | ···················· | ···················· | ···················· | ···················· | |
| E | 9076 | hCoV-19/USA/WI-UW-589/2020\|EPI_ISL_484964\|2020-06- | ···················· | ···················· | ···················· | ···················· | ···················· | ···················· | |
| E | 9077 | hCoV-19/USA/WI-UW-590/2020\|EPI_ISL_484965\|2020-06- | ···················· | ···················· | ···················· | ···················· | ···················· | ···················· | |
| E | 9078 | hCoV-19/USA/WI-UW-591/2020\|EPI_ISL_484966\|2020-06- | ···················· | ···················· | ···················· | ···················· | ···················· | ···················· | |
| E | 9079 | hCoV-19/USA/IL-UW-592/2020\|EPI_ISL_484967\|2020-06- | ···················· | ···················· | ···················· | ···················· | ···················· | ···················· | |
| E | 9080 | hCoV-19/USA/WI-UW-594/2020\|EPI_ISL_484969\|2020-06- | ···················· | ···················· | ···················· | ···················· | ···················· | ···················· | |
| E | 9081 | hCoV-19/USA/WA-UW-2565/2020\|EPI_ISL_486000\|2020-03 | ···················· | ···················· | ···················· | ···················· | ···················· | ···················· | |
| E | 9082 | hCoV-19/USA/WA-UW-2566/2020\|EPI_ISL_486001\|2020-03 | ···················· | ···················· | ···················· | ···················· | ···················· | ···················· | |
| E | 9083 | hCoV-19/USA/WA-UW-2572/2020\|EPI_ISL_486002\|2020-03 | ···················· | ···················· | ···················· | ···················· | ···················· | ···················· | |
| E | 9084 | hCoV-19/USA/WA-UW-2599/2020\|EPI_ISL_486003\|2020-03 | ···················· | ···················· | ···················· | ···················· | ···················· | ···················· | |
| E | 9085 | hCoV-19/USA/WA-UW-2663/2020\|EPI_ISL_486011\|2020-03 | ···················· | ···················· | ···················· | ···················· | ···················· | ···················· | |
| E | 9086 | hCoV-19/USA/WA-UW-2665/2020\|EPI_ISL_486012\|2020-03 | ···················· | ···················· | ···················· | ···················· | ···················· | ···················· | |
| E | 9087 | hCoV-19/USA/WA-UW-2670/2020\|EPI_ISL_486013\|2020-03 | ···················· | ···················· | ···················· | ···················· | ···················· | ···················· | |
| E | 9088 | hCoV-19/USA/WA-UW-2679/2020\|EPI_ISL_486014\|2020-04 | ···················· | ···················· | ···················· | ···················· | ···················· | ···················· | |
| E | 9089 | hCoV-19/USA/WA-UW-2805/2020\|EPI_ISL_486037\|2020-03 | ···················· | ···················· | ···················· | ···················· | ···················· | ···················· | |
| E | 9090 | hCoV-19/USA/WA-UW-2818/2020\|EPI_ISL_486039\|2020-03 | ···················· | ···················· | ···················· | ···················· | ···················· | ···················· | |
| E | 9091 | hCoV-19/USA/WA-UW-2862/2020\|EPI_ISL_486040\|2020-03 | ···················· | ···················· | ···················· | ···················· | ···················· | ···················· | |
| E | 9092 | hCoV-19/USA/WA-UW-2872/2020\|EPI_ISL_486041\|2020-03 | ···················· | ···················· | ···················· | ···················· | ···················· | ···················· | |
| E | 9093 | hCoV-19/USA/WA-UW-2875/2020\|EPI_ISL_486042\|2020-03 | ···················· | ···················· | ···················· | ···················· | ···················· | ···················· | |
| E | 9094 | hCoV-19/USA/WA-UW-2877/2020\|EPI_ISL_486043\|2020-03 | ···················· | ···················· | ···················· | ···················· | ···················· | ···················· | |
| E | 9095 | hCoV-19/USA/WA-UW-2879/2020\|EPI_ISL_486044\|2020-03 | ···················· | ···················· | ···················· | ···················· | ···················· | ···················· | |
| E | 9096 | hCoV-19/USA/WA-UW-2902/2020\|EPI_ISL_486046\|2020-03 | ···················· | ···················· | ···················· | ···················· | ···················· | ···················· | |
| E | 9097 | hCoV-19/USA/WA-UW-2909/2020\|EPI_ISL_486047\|2020-03 | ···················· | ···················· | ···················· | ···················· | ···················· | ···················· | |
| E | 9098 | hCoV-19/USA/WA-UW-2910/2020\|EPI_ISL_486048\|2020-03 | ···················· | ···················· | ···················· | ···················· | ···················· | ···················· | |
| E | 9099 | hCoV-19/USA/WA-UW-2911/2020\|EPI_ISL_486049\|2020-03 | ···················· | ···················· | ···················· | ···················· | ···················· | ···················· | |
| E | 9100 | hCoV-19/USA/WA-UW-2916/2020\|EPI_ISL_486050\|2020-03 | ···················· | ···················· | ···················· | ···················· | ···················· | ···················· | |
| E | 9101 | hCoV-19/USA/WA-UW-2921/2020\|EPI_ISL_486051\|2020-03 | ···················· | ···················· | ···················· | ···················· | ···················· | ···················· | |
| E | 9102 | hCoV-19/USA/WA-UW-2928/2020\|EPI_ISL_486052\|2020-03 | ···················· | ···················· | ···················· | ···················· | ···················· | ··················C· | 1 |
| E | 9103 | hCoV-19/USA/WA-UW-2929/2020\|EPI_ISL_486053\|2020-03 | ···················· | ···················· | ···················· | ···················· | ···················· | ···················· | |
| E | 9104 | hCoV-19/USA/WA-UW-2935/2020\|EPI_ISL_486054\|2020-03 | ···················· | ···················· | ···················· | ···················· | ···················· | ···················· | |
| E | 9105 | hCoV-19/USA/WA-UW-2936/2020\|EPI_ISL_486055\|2020-03 | ···················· | ···················· | ···················· | ···················· | ···················· | ···················· | |
| E | 9106 | hCoV-19/USA/WA-UW-2937/2020\|EPI_ISL_486056\|2020-03 | ···················· | ···················· | ···················· | ···················· | ···················· | ···················· | |
| E | 9107 | hCoV-19/USA/WA-UW-2940/2020\|EPI_ISL_486057\|2020-03 | ···················· | ···················· | ···················· | ···················· | ···················· | ···················· | |
| E | 9108 | hCoV-19/USA/WA-UW-2943/2020\|EPI_ISL_486058\|2020-03 | ···················· | ···················· | ···················· | ···················· | ···················· | ···················· | |
| E | 9109 | hCoV-19/USA/WA-UW-2953/2020\|EPI_ISL_486059\|2020-03 | ···················· | ···················· | ···················· | ···················· | ···················· | ···················· | |
| E | 9110 | hCoV-19/USA/WA-UW-2960/2020\|EPI_ISL_486060\|2020-03 | ···················· | ···················· | ···················· | ···················· | ···················· | ···················· | |
| E | 9111 | hCoV-19/USA/WA-UW-2961/2020\|EPI_ISL_486061\|2020-03 | ···················· | ···················· | ···················· | ···················· | ···················· | ···················· | |
| E | 9112 | hCoV-19/USA/WA-UW-2963/2020\|EPI_ISL_486062\|2020-03 | ···················· | ···················· | ···················· | ···················· | ···················· | ···················· | |
| E | 9113 | hCoV-19/USA/WA-UW-2964/2020\|EPI_ISL_486063\|2020-03 | ···················· | ···················· | ···················· | ···················· | ···················· | ···················· | |
| E | 9114 | hCoV-19/USA/WA-UW-2971/2020\|EPI_ISL_486064\|2020-03 | ···················· | ···················· | ···················· | ···················· | ···················· | ···················· | |

E 9115 hCoV-19/USA/WA-UW-2976/2020|EPI_ISL_486065|2020-03 ···················· ···················· ····················· ···················· ···················· ····················
E 9116 hCoV-19/USA/WA-UW-2980/2020|EPI_ISL_486066|2020-03 ···················· ···················· ····················· ···················· ···················· ····················
E 9117 hCoV-19/USA/WA-UW-2982/2020|EPI_ISL_486067|2020-03 ···················· ···················· ····················· ···················· ···················· ····················
E 9118 hCoV-19/USA/WA-UW-2983/2020|EPI_ISL_486068|2020-03 ···················· ···················· ····················· ···················· ···················· ····················
E 9119 hCoV-19/USA/WA-UW-2986/2020|EPI_ISL_486069|2020-03 ···················· ···················· ····················· ···················· ···················· ····················
E 9120 hCoV-19/USA/WA-UW-2987/2020|EPI_ISL_486070|2020-03 ···················· ···················· ····················· ···················· ···················· ····················
E 9121 hCoV-19/USA/WA-UW-2997/2020|EPI_ISL_486071|2020-03 ···················· ···················· ····················· ···················· ···················· ····················
E 9122 hCoV-19/USA/WA-UW-3001/2020|EPI_ISL_486072|2020-03 ···················· ···················· ····················· ···················· ···················· ····················
E 9123 hCoV-19/USA/WA-UW-3003/2020|EPI_ISL_486074|2020-03 ···················· ···················· ····················· ···················· ···················· ····················
E 9124 hCoV-19/USA/WA-UW-3007/2020|EPI_ISL_486075|2020-03 ···················· ···················· ····················· ···················· ···················· ····················
E 9125 hCoV-19/USA/WA-UW-3008/2020|EPI_ISL_486076|2020-03 ···················· ···················· ····················· ···················· ···················· ····················
E 9126 hCoV-19/USA/WA-UW-3009/2020|EPI_ISL_486077|2020-03 ···················· ···················· ····················· ···················· ···················· ····················
E 9127 hCoV-19/USA/WA-UW-3012/2020|EPI_ISL_486078|2020-03 ···················· ···················· ····················· ···················· ···················· ····················
E 9128 hCoV-19/USA/WA-UW-3018/2020|EPI_ISL_486079|2020-03 ···················· ···················· ····················· ···················· ···················· ····················
E 9129 hCoV-19/DRC/4516/2020|EPI_ISL_487192|2020-04-28 ···················· ···················· ····················· ···················· ···················· ····················
E 9130 hCoV-19/USA/UT-02538/2020|EPI_ISL_487199|2020-04-2 ···················· ···················· ····················· ···················· ···················· ····················
E 9131 hCoV-19/South Africa/KRISP-0373/2020|EPI_ISL_48732 ···················· ···················· ····················· ···················· ···················· ····················
E 9132 hCoV-19/South Africa/KRISP-0375/2020|EPI_ISL_48732 ···················· ···················· ····················· ···················· ···················· ····················
E 9133 hCoV-19/South Africa/KRISP-0380/2020|EPI_ISL_48732 ···················· ···················· ····················· ···················· ···················· ····················
E 9134 hCoV-19/South Africa/KRISP-0386/2020|EPI_ISL_48732 ···················· ···················· ····················· ···················· ···················· ····················
E 9135 hCoV-19/South Africa/KRISP-0387/2020|EPI_ISL_48733 ···················· ···················· ····················· ···················· ···················· ····················
E 9136 hCoV-19/South Africa/KRISP-0389/2020|EPI_ISL_48733 ···················· ···················· ····················· ···················· ···················· ····················
E 9137 hCoV-19/South Africa/KRISP-0391/2020|EPI_ISL_48733 ···················· ···················· ····················· ···················· ···················· ····················
E 9138 hCoV-19/Bangladesh/BCSIR-NILMRC-229/2020|EPI_ISL_4 ···················· ···················· ····················· ···················· ···················· ····················
E 9139 hCoV-19/Bangladesh/BCSIR-NILMRC-230/2020|EPI_ISL_4 ···················· ···················· ····················· ···················· ···················· ····················
E 9140 hCoV-19/Bangladesh/BCSIR-NILMRC-231/2020|EPI_ISL_4 ···················· ···················· ····················· ···················· ···················· ····················
E 9141 hCoV-19/Bangladesh/BCSIR-NILMRC-232/2020|EPI_ISL_4 ···················· ·A·················· ····················· ···················· ···················· ···A················
E 9142 hCoV-19/Bangladesh/BCSIR-NILMRC-233/2020|EPI_ISL_4 ···················· ···················· ····················· ···················· ···················· ····················
E 9143 hCoV-19/Greece/8594_HPI/2020|EPI_ISL_487377|2020-0 ···················· ···················· ····················· ···················· ···················· ····················
E 9144 hCoV-19/Bangladesh/BCSIR-NILMRC-234/2020|EPI_ISL_4 ···················· ···················· ····················· ···················· ···················· ····················
E 9145 hCoV-19/Bangladesh/BCSIR-NILMRC-235/2020|EPI_ISL_4 ···················· ···················· ····················· ···················· ···················· ····················
E 9146 hCoV-19/Greece/9060_HPI/2020|EPI_ISL_487381|2020-0 ···················· ···················· ····················· ···················· ···················· ····················
E 9147 hCoV-19/Bangladesh/BCSIR-NILMRC-242/2020|EPI_ISL_4 ···················· ···················· ····················· ···················· ···················· ····················
E 9148 hCoV-19/Bangladesh/BCSIR-NILMRC-243/2020|EPI_ISL_4 ···················· ···················· ····················· ···················· ···················· ····················
E 9149 hCoV-19/Bangladesh/BCSIR-NILMRC-244/2020|EPI_ISL_4 ···················· ···················· ····················· ···················· ···················· ····················
E 9150 hCoV-19/Bangladesh/BCSIR-NILMRC-245/2020|EPI_ISL_4 ···················· ···················· ····················· ···················· ···················· ····················
E 9151 hCoV-19/Bangladesh/BCSIR-NILMRC-236/2020|EPI_ISL_4 ···················· ···················· ····················· ···················· ···················· ····················
E 9152 hCoV-19/Bangladesh/BCSIR-NILMRC-237/2020|EPI_ISL_4 ···················· ···················· ····················· ···················· ···················· ····················
E 9153 hCoV-19/Bangladesh/BCSIR-NILMRC-238/2020|EPI_ISL_4 ···················· ···················· ····················· ···················· ···················· ····················
E 9154 hCoV-19/Bangladesh/BCSIR-NILMRC-239/2020|EPI_ISL_4 ···················· ···················· ····················· ···················· ···················· ····················
E 9155 hCoV-19/Bangladesh/BCSIR-NILMRC-240/2020|EPI_ISL_4 ···················· ···················· ····················· ···················· ···················· ····················
E 9156 hCoV-19/Bangladesh/BCSIR-NILMRC-241/2020|EPI_ISL_4 ···················· ···················· ····················· ···················· ···················· ····················
E 9157 hCoV-19/Germany/NW-MPP-10/2020|EPI_ISL_487398|2020 ···················· ···················· ····················· ···················· ···················· ····················
E 9158 hCoV-19/Germany/NW-MPP-11/2020|EPI_ISL_487399|2020 ···················· ···················· ····················· ···················· ···················· ····················
E 9159 hCoV-19/England/NORT-283267/2020|EPI_ISL_488655|20 ···················· ···················· ····················· ···················· ···················· ····················
E 9160 hCoV-19/England/NORT-295D16/2020|EPI_ISL_488656|20 ···················· ···················· ····················· ···················· ···················· ····················
E 9161 hCoV-19/England/NORT-284A13/2020|EPI_ISL_488657|20 ···················· ···················· ····················· ···················· ···················· ····················
E 9162 hCoV-19/England/NORT-28472B/2020|EPI_ISL_488658|20 ···················· ···················· ····················· ···················· ···················· ····················
E 9163 hCoV-19/England/NORT-282FE9/2020|EPI_ISL_488659|20 ···················· ···················· ····················· ···················· ···················· ····················
E 9164 hCoV-19/England/NORT-283197/2020|EPI_ISL_488660|20 ···················· ···················· ····················· ···················· ···················· ····················
E 9165 hCoV-19/England/NORT-283DC0/2020|EPI_ISL_488661|20 ···················· ···················· ····················· ···················· ···················· ····················
E 9166 hCoV-19/England/NORT-282972/2020|EPI_ISL_488662|20 ···················· ···················· ····················· ···················· ···················· ····················
E 9167 hCoV-19/England/NORT-283865/2020|EPI_ISL_488663|20 ···················· ···················· ····················· ···················· ···················· ····················
E 9168 hCoV-19/England/NORT-284C77/2020|EPI_ISL_488664|20 ···················· ···················· ····················· ···················· ···················· ····················
E 9169 hCoV-19/England/NORT-295332/2020|EPI_ISL_488665|20 ···················· ···················· ····················· ···················· ···················· ····················
E 9170 hCoV-19/England/NORT-2843AF/2020|EPI_ISL_488673|20 ···················· ···················· ····················· ···················· ···················· ····················
E 9171 hCoV-19/England/NORT-284460/2020|EPI_ISL_488674|20 ···················· ···················· ····················· ···················· ···················· ····················
E 9172 hCoV-19/England/NORT-284B89/2020|EPI_ISL_488675|20 ···················· ···················· ····················· ···················· ···················· ····················
E 9173 hCoV-19/England/NORT-295D9E/2020|EPI_ISL_488676|20 ···················· ···················· ····················· ···················· ···················· ····················
E 9174 hCoV-19/England/NORT-295921/2020|EPI_ISL_488699|20 ···················· ···················· ····················· ···················· ···················· ····················
E 9175 hCoV-19/Switzerland/VD-ETHZ-180052/2020|EPI_ISL_48 ···················· ·A·················· ····················· ···················· ···················· ····················
E 9176 hCoV-19/Peru/LIM-INS-009/2020|EPI_ISL_489987|2020- ···················· ···················· ····················· ···················· ···················· ····················
E 9177 hCoV-19/Peru/LIM-INS-011/2020|EPI_ISL_489988|2020- ···················· ···················· ····················· ···················· ···················· ····················
E 9178 hCoV-19/Peru/ICA-INS-012/2020|EPI_ISL_489989|2020- ···················· ···················· ····················· ···················· ···················· ····················
E 9179 hCoV-19/Peru/LIM-INS-014/2020|EPI_ISL_489990|2020- ···················· ···················· ····················· ···················· ···················· ····················
E 9180 hCoV-19/Thailand/Samut Songkhram-0942/2020|EPI_ISL ···················· ··········T········· ····················· ···················· ···················· ····················
E 9181 hCoV-19/Malaysia/IMR-WC9205/2020|EPI_ISL_489992|20 ···················· ···················· ····················· ···················· ···················· ····················
E 9182 hCoV-19/Malaysia/IMR-WC9185/2020|EPI_ISL_489994|20 ···················· ···················· ····················· ···················· ···················· ····················
E 9183 hCoV-19/India/UN-UN3/2020|EPI_ISL_489995|2020 ···················· ···················· ····················· ···················· ····N····N·········· ···N············N···
E 9184 hCoV-19/Saudi Arabia/477/2020|EPI_ISL_489996|2020- ···················· ···················· ····················· ···················· ···················· ····················
E 9185 hCoV-19/Peru/LIM-INS-018/2020|EPI_ISL_490975|2020- ···················· ···················· ····················· ···················· ···················· ····················
E 9186 hCoV-19/Peru/LIM-INS-020/2020|EPI_ISL_490976|2020- ···················· ···················· ····················· ···················· ···················· ····················
E 9187 hCoV-19/USA/CT-UW-3123/2020|EPI_ISL_490980|2020-03 ···················· ···················· ····················· ···················· ···················· ····················
E 9188 hCoV-19/USA/CT-UW-3417/2020|EPI_ISL_490982|2020-03 ···················· ···················· ····················· ···················· ···················· ····················
E 9189 hCoV-19/USA/CT-UW-3423/2020|EPI_ISL_490983|2020-03 ···················· ···················· ····················· ···················· ···················· ····················
E 9190 hCoV-19/USA/CT-UW-3425/2020|EPI_ISL_490984|2020-03 ···················· ···················· ····················· ···················· ···················· ····················
E 9191 hCoV-19/USA/CT-UW-3450/2020|EPI_ISL_490985|2020-03 ···················· ···················· ····················· ···················· ···················· ····················
E 9192 hCoV-19/USA/CT-UW-3465/2020|EPI_ISL_490986|2020-03 ···················· ···················· ····················· ···················· ···················· ····················
E 9193 hCoV-19/USA/CT-UW-3470/2020|EPI_ISL_490987|2020-03 ···················· ···················· ····················· ···················· ···················· ····················
E 9194 hCoV-19/USA/FL-UW-MAYO68/2020|EPI_ISL_490988|2020- ···················· ···················· ····················· ···················· ···················· ····················

| | | | | | | | | |
|---|---|---|---|---|---|---|---|---|
| E | 9195 | hCoV-19/USA/ID-UW-3172/2020\|EPI_ISL_490989\|2020-03 | ···················· | ···················· | ···················· | ···················· | ···················· | ···················· |
| E | 9196 | hCoV-19/USA/MN-MDH-1301/2020\|EPI_ISL_495600\|2020-0 | ···················· | ···················· | ···················· | ···················· | ···················· | ···················· |
| E | 9197 | hCoV-19/USA/MN-MDH-1302/2020\|EPI_ISL_495601\|2020-0 | ···················· | ···················· | ···················· | ···················· | ···················· | ···················· |
| E | 9198 | hCoV-19/USA/MN-MDH-1303/2020\|EPI_ISL_495602\|2020-0 | ···················· | ···················· | ···················· | ···················· | ···················· | ···················· |
| E | 9199 | hCoV-19/USA/MN-MDH-1304/2020\|EPI_ISL_495603\|2020-0 | ···················· | ···················· | ···················· | ···················· | ···················· | ···················· |
| E | 9200 | hCoV-19/USA/MN-MDH-1305/2020\|EPI_ISL_495604\|2020-0 | ···················· | ···················· | ···················· | ···················· | ···················· | ···················· |
| E | 9201 | hCoV-19/USA/MN-MDH-1306/2020\|EPI_ISL_495605\|2020-0 | ···················· | ···················· | ···················· | ···················· | ···················· | ···················· |
| E | 9202 | hCoV-19/USA/MN-MDH-1307/2020\|EPI_ISL_495606\|2020-0 | ···················· | ···················· | ···················· | ···················· | ···················· | ···················· |
| E | 9203 | hCoV-19/USA/MN-MDH-1308/2020\|EPI_ISL_495607\|2020-0 | ···················· | ···················· | ···················· | ···················· | ···················· | ···················· |
| E | 9204 | hCoV-19/USA/MN-MDH-1309/2020\|EPI_ISL_495608\|2020-0 | ···················· | ···················· | ···················· | ···················· | ···················· | ···················· |
| E | 9205 | hCoV-19/USA/MN-MDH-1310/2020\|EPI_ISL_495609\|2020-0 | ···················· | ···················· | ···················· | ···················· | ···················· | ···················· |
| E | 9206 | hCoV-19/USA/MN-MDH-1311/2020\|EPI_ISL_495610\|2020-0 | ···················· | ···················· | ···················· | ···················· | ···················· | ···················· |
| E | 9207 | hCoV-19/USA/MN-MDH-1312/2020\|EPI_ISL_495611\|2020-0 | ···················· | ····G··············· | ···················· | ···················· | ···················· | ···················· |
| E | 9208 | hCoV-19/USA/MN-MDH-1313/2020\|EPI_ISL_495612\|2020-0 | ···················· | ···················· | ···················· | ···················· | ···················· | ···················· |
| E | 9209 | hCoV-19/USA/MN-MDH-1314/2020\|EPI_ISL_495613\|2020-0 | ···················· | ···················· | ···················· | ···················· | ···················· | ···················· |
| E | 9210 | hCoV-19/USA/MN-MDH-1315/2020\|EPI_ISL_495614\|2020-0 | ···················· | ···················· | ···················· | ···················· | ···················· | ···················· |
| E | 9211 | hCoV-19/USA/MN-MDH-1316/2020\|EPI_ISL_495615\|2020-0 | ···················· | ···················· | ···················· | ···················· | ···················· | ···················· |
| E | 9212 | hCoV-19/USA/MN-MDH-1317/2020\|EPI_ISL_495616\|2020-0 | ···················· | ···················· | ···················· | ···················· | ···················· | ···················· |
| E | 9213 | hCoV-19/USA/MN-MDH-1318/2020\|EPI_ISL_495617\|2020-0 | ···················· | ···················· | ···················· | ···················· | ···················· | ···················· |
| E | 9214 | hCoV-19/USA/MN-MDH-1319/2020\|EPI_ISL_495618\|2020-0 | ···················· | ···················· | ···················· | ···················· | ···················· | ···················· |
| E | 9215 | hCoV-19/USA/MN-MDH-1320/2020\|EPI_ISL_495619\|2020-0 | ···················· | ···················· | ···················· | ···················· | ···················· | ···················· |
| E | 9216 | hCoV-19/USA/WA-S1475/2020\|EPI_ISL_495650\|2020-05-1 | ···················· | ···················· | ···················· | ···················· | ···················· | ···················· |
| E | 9217 | hCoV-19/India/OR-ILSCV34939/2020\|EPI_ISL_481186\|20 | ···················· | ···················· | ···················· | ···················· | ···················· | ···················· |
| E | 9218 | hCoV-19/India/OR-ILSCV35141/2020\|EPI_ISL_481187\|20 | ···················· | ···················· | ···················· | ···················· | ···················· | ···················· |
| E | 9219 | hCoV-19/India/OR-ILSCV35317/2020\|EPI_ISL_481189\|20 | ···················· | ···················· | ···················· | ···················· | ···················· | ···················· |
| E | 9220 | hCoV-19/India/OR-ILSCV35333/2020\|EPI_ISL_481190\|20 | ···················· | ···················· | ···················· | ···················· | ···················· | ···················· |
| E | 9221 | hCoV-19/India/OR-ILSCV35350/2020\|EPI_ISL_481191\|20 | ···················· | ···················· | ···················· | ···················· | ···················· | ···················· |
| E | 9222 | hCoV-19/India/OR-ILSCV35351/2020\|EPI_ISL_481192\|20 | ···················· | ···················· | ···················· | ···················· | ···················· | ···················· |
| E | 9223 | hCoV-19/India/OR-ILSCV35353/2020\|EPI_ISL_481193\|20 | ···················· | ···················· | ···················· | ···················· | ···················· | ···················· |
| E | 9224 | hCoV-19/India/OR-ILSCV35366/2020\|EPI_ISL_481194\|20 | ···················· | ···················· | ···················· | ···················· | ···················· | ···················· |
| E | 9225 | hCoV-19/India/OR-ILSCV35376/2020\|EPI_ISL_481195\|20 | ···················· | ···················· | ···················· | ···················· | ···················· | ···················· |
| E | 9226 | hCoV-19/India/OR-ILSCV35456/2020\|EPI_ISL_481196\|20 | ···················· | ···················· | ···················· | ···················· | ···················· | ···················· |
| E | 9227 | hCoV-19/India/OR-ILSCV35457/2020\|EPI_ISL_481197\|20 | ···················· | ···················· | ···················· | ···················· | ···················· | ···················· |
| E | 9228 | hCoV-19/India/OR-ILSCV35461/2020\|EPI_ISL_481198\|20 | ···················· | ···················· | ···················· | ···················· | ···················· | ···················· |
| E | 9229 | hCoV-19/India/OR-ILSCV35475/2020\|EPI_ISL_481199\|20 | ···················· | ···················· | ···················· | ···················· | ···················· | ···················· |
| E | 9230 | hCoV-19/USA/WA-UW-2606/2020\|EPI_ISL_486004\|2020-03 | ···················· | ···················· | ···················· | ···················· | ···················· | ···················· |
| E | 9231 | hCoV-19/USA/WA-UW-2608/2020\|EPI_ISL_486005\|2020-03 | ···················· | ···················· | ···················· | ···················· | ···················· | ···················· |
| E | 9232 | hCoV-19/USA/WA-UW-2609/2020\|EPI_ISL_486006\|2020-03 | ···················· | ···················· | ···················· | ···················· | ···················· | ···················· |
| E | 9233 | hCoV-19/USA/WA-UW-2618/2020\|EPI_ISL_486008\|2020-03 | ···················· | ···················· | ···················· | ···················· | ···················· | ···················· |
| E | 9234 | hCoV-19/USA/WA-UW-2627/2020\|EPI_ISL_486009\|2020-03 | ···················· | ···················· | ···················· | ···················· | ···················· | ···················· |
| E | 9235 | hCoV-19/USA/WA-UW-2651/2020\|EPI_ISL_486010\|2020-03 | ···················· | ···················· | ···················· | ···················· | ···················· | ···················· |
| E | 9236 | hCoV-19/USA/WA-UW-2686/2020\|EPI_ISL_486015\|2020-04 | ···················· | ···················· | ···················· | ···················· | ···················· | ···················· |
| E | 9237 | hCoV-19/USA/WA-UW-2705/2020\|EPI_ISL_486017\|2020-03 | ···················· | ···················· | ···················· | ···················· | ···················· | ···················· |
| E | 9238 | hCoV-19/USA/WA-UW-2725/2020\|EPI_ISL_486018\|2020-03 | ···················· | ···················· | ···················· | ···················· | ···················· | ···················· |
| E | 9239 | hCoV-19/USA/WA-UW-2729/2020\|EPI_ISL_486019\|2020-04 | ···················· | ···················· | ···················· | ···················· | ···················· | ···················· |
| E | 9240 | hCoV-19/USA/WA-UW-2730/2020\|EPI_ISL_486020\|2020-03 | ···················· | ···················· | ···················· | ···················· | ···················· | ···················· |
| E | 9241 | hCoV-19/USA/WA-UW-2732/2020\|EPI_ISL_486021\|2020-03 | ···················· | ···················· | ···················· | ···················· | ···················· | ···················· |
| E | 9242 | hCoV-19/USA/WA-UW-2735/2020\|EPI_ISL_486022\|2020-04 | ···················· | ···················· | ···················· | ···················· | ···················· | ···················· |
| E | 9243 | hCoV-19/USA/WA-UW-2736/2020\|EPI_ISL_486023\|2020-04 | ···················· | ···················· | ···················· | ···················· | ···················· | ···················· |
| E | 9244 | hCoV-19/USA/WA-UW-2737/2020\|EPI_ISL_486024\|2020-04 | ···················· | ···················· | ···················· | ···················· | ···················· | ···················· |
| E | 9245 | hCoV-19/USA/WA-UW-2743/2020\|EPI_ISL_486025\|2020-04 | ···················· | ···················· | ···················· | ···················· | ···················· | ···················· |
| E | 9246 | hCoV-19/USA/WA-UW-2744/2020\|EPI_ISL_486026\|2020-04 | ···················· | ···················· | ···················· | ···················· | ···················· | ···················· |
| E | 9247 | hCoV-19/USA/WA-UW-2747/2020\|EPI_ISL_486027\|2020-04 | ···················· | ···················· | ···················· | ···················· | ···················· | ···················· |
| E | 9248 | hCoV-19/USA/WA-UW-2756/2020\|EPI_ISL_486028\|2020-03 | ···················· | ···················· | ···················· | ···················· | ···················· | ···················· |
| E | 9249 | hCoV-19/USA/WA-UW-2761/2020\|EPI_ISL_486029\|2020-04 | ···················· | ···················· | ···················· | ···················· | ···················· | ···················· |
| E | 9250 | hCoV-19/USA/WA-UW-2767/2020\|EPI_ISL_486030\|2020-03 | ···················· | ···················· | ···················· | ···················· | ···················· | ···················· |
| E | 9251 | hCoV-19/USA/WA-UW-2776/2020\|EPI_ISL_486032\|2020-04 | ···················· | ···················· | ···················· | ···················· | ···················· | ···················· |
| E | 9252 | hCoV-19/USA/WA-UW-2777/2020\|EPI_ISL_486033\|2020-03 | ···················· | ···················· | ···················· | ···················· | ···················· | ···················· |
| E | 9253 | hCoV-19/USA/WA-UW-2778/2020\|EPI_ISL_486034\|2020-03 | ···················· | ···················· | ···················· | ···················· | ···················· | ···················· |
| E | 9254 | hCoV-19/USA/WA-UW-2781/2020\|EPI_ISL_486035\|2020-03 | ···················· | ···················· | ···················· | ···················· | ···················· | ···················· |
| E | 9255 | hCoV-19/USA/WA-UW-2795/2020\|EPI_ISL_486036\|2020-04 | ···················· | ···················· | ···················· | ···················· | ···················· | ···················· |
| E | 9256 | hCoV-19/South Africa/KRISP-0336/2020\|EPI_ISL_48730 | ···················· | ···················· | ···················· | ···················· | ···················· | ···················· |
| E | 9257 | hCoV-19/South Africa/KRISP-0347/2020\|EPI_ISL_48730 | ···················· | ···················· | ···················· | ···················· | ···················· | ···················· |
| E | 9258 | hCoV-19/South Africa/KRISP-0350/2020\|EPI_ISL_48731 | ···················· | ···················· | ···················· | ···················· | ···················· | ···················· |
| E | 9259 | hCoV-19/South Africa/KRISP-0352/2020\|EPI_ISL_48731 | ···················· | ···················· | ···················· | ···················· | ···················· | ···················· |
| E | 9260 | hCoV-19/South Africa/KRISP-0356/2020\|EPI_ISL_48731 | ···················· | ···················· | ···················· | ···················· | ···················· | ···················· |
| E | 9261 | hCoV-19/South Africa/KRISP-0360/2020\|EPI_ISL_48731 | ···················· | ···················· | ···················· | ···················· | ···················· | ···················· |
| E | 9262 | hCoV-19/South Africa/KRISP-0365/2020\|EPI_ISL_48731 | ···················· | ···················· | ···················· | ···················· | ···················· | ···················· |
| E | 9263 | hCoV-19/South Africa/KRISP-0367/2020\|EPI_ISL_48731 | ···················· | ···················· | ···················· | ···················· | ···················· | ···················· |
| E | 9264 | hCoV-19/South Africa/KRISP-0368/2020\|EPI_ISL_48731 | ···················· | ···················· | ···················· | ···················· | ···················· | ···················· |
| E | 9265 | hCoV-19/South Africa/KRISP-0369/2020\|EPI_ISL_48732 | ···················· | ···················· | ···················· | ···················· | ···················· | ···················· |
| E | 9266 | hCoV-19/South Africa/KRISP-0370/2020\|EPI_ISL_48732 | ···················· | ···················· | ···················· | ···················· | ···················· | ···················· |
| E | 9267 | hCoV-19/South Africa/KRISP-0371/2020\|EPI_ISL_48732 | ···················· | ···················· | ···················· | ···················· | ···················· | ···················· |
| E | 9268 | hCoV-19/South Africa/KRISP-0393/2020\|EPI_ISL_48733 | ···················· | ···················· | ···················· | ···················· | ···················· | ···················· |
| E | 9269 | hCoV-19/South Africa/KRISP-0394/2020\|EPI_ISL_48733 | ···················· | ···················· | ···················· | ···················· | ···················· | ···················· |
| E | 9270 | hCoV-19/South Africa/KRISP-0395/2020\|EPI_ISL_48733 | ···················· | ···················· | ···················· | ···················· | ···················· | ···················· |
| E | 9271 | hCoV-19/South Africa/KRISP-0396/2020\|EPI_ISL_48733 | ···················· | ···················· | ···················· | ···················· | ···················· | ···················· |
| E | 9272 | hCoV-19/South Africa/KRISP-0397/2020\|EPI_ISL_48734 | ···················· | ···················· | ···················· | ···················· | ···················· | ···················· |
| E | 9273 | hCoV-19/Bangladesh/BCSIR-NILMRC-222/2020\|EPI_ISL_4 | ···················· | ···················· | ···················· | ···················· | ···················· | ···A················ |
| E | 9274 | hCoV-19/Bangladesh/BCSIR-NILMRC-223/2020\|EPI_ISL_4 | ···················· | ···················· | ···················· | ···················· | ···················· | ···················· |

| | | | | | | | | |
|---|---|---|---|---|---|---|---|---|
| E | 9275 | hCoV-19/Bangladesh/BCSIR-NILMRC-224/2020\|EPI_ISL_4 | ···················· | ···················· | ···················· | ···················· | ···················· | ···················· |
| E | 9276 | hCoV-19/Bangladesh/BCSIR-NILMRC-226/2020\|EPI_ISL_4 | ···················· | ···················· | ···················· | ···················· | ···················· | ···················· |
| E | 9277 | hCoV-19/Bangladesh/BCSIR-NILMRC-227/2020\|EPI_ISL_4 | ···················· | ···················· | ···················· | ···················· | ···················· | ···················· |
| E | 9278 | hCoV-19/England/NORT-282F43/2020\|EPI_ISL_488601\|20 | ···················· | ············A······· | ···················· | ···················· | ···················· | ···················· |
| E | 9279 | hCoV-19/England/NORT-29552D/2020\|EPI_ISL_488604\|20 | ···················· | ···················· | ···················· | ···················· | ···················· | ···················· |
| E | 9280 | hCoV-19/England/NORT-283294/2020\|EPI_ISL_488605\|20 | ···················· | ···················· | ···················· | ···················· | ···················· | ···················· |
| E | 9281 | hCoV-19/England/NORT-294BA4/2020\|EPI_ISL_488606\|20 | ···················· | ···················· | ···················· | ···················· | ···················· | ···················· |
| E | 9282 | hCoV-19/England/NORT-295499/2020\|EPI_ISL_488607\|20 | ···················· | ···················· | ···················· | ···················· | ···················· | ···················· |
| E | 9283 | hCoV-19/England/NORT-295B3A/2020\|EPI_ISL_488610\|20 | ···················· | ···················· | ···················· | ···················· | ···················· | ···················· |
| E | 9284 | hCoV-19/England/NORT-2846F1/2020\|EPI_ISL_488611\|20 | ···················· | ···················· | ···················· | ···················· | ···················· | ···················· |
| E | 9285 | hCoV-19/England/NORT-283D93/2020\|EPI_ISL_488612\|20 | ···················· | ···················· | ···················· | ···················· | ···················· | ···················· |
| E | 9286 | hCoV-19/England/NORT-295208/2020\|EPI_ISL_488613\|20 | ···················· | ···················· | ···················· | ···················· | ···················· | ···················· |
| E | 9287 | hCoV-19/England/NORT-282796/2020\|EPI_ISL_488614\|20 | ···················· | ···················· | ···················· | ···················· | ···················· | ···················· |
| E | 9288 | hCoV-19/England/NORT-283601/2020\|EPI_ISL_488615\|20 | ···················· | ···················· | ···················· | ···················· | ···················· | ···················· |
| E | 9289 | hCoV-19/England/NORT-2957BE/2020\|EPI_ISL_488616\|20 | ···················· | ···················· | ···················· | ···················· | ···················· | ···················· |
| E | 9290 | hCoV-19/England/NORT-282D85/2020\|EPI_ISL_488617\|20 | ···················· | ···················· | ···················· | ···················· | ···················· | ···················· |
| E | 9291 | hCoV-19/England/NORT-2958BB/2020\|EPI_ISL_488618\|20 | ···················· | ···················· | ···················· | ···················· | ···················· | ···················· |
| E | 9292 | hCoV-19/England/NORT-295253/2020\|EPI_ISL_488619\|20 | ···················· | ···················· | ···················· | ···················· | ···················· | ···················· |
| E | 9293 | hCoV-19/England/NORT-282550/2020\|EPI_ISL_488620\|20 | ···················· | ···················· | ···················· | ···················· | ···················· | ···················· |
| E | 9294 | hCoV-19/England/NORT-283B7B/2020\|EPI_ISL_488621\|20 | ···················· | ···················· | ···················· | ···················· | ···················· | ···················· |
| E | 9295 | hCoV-19/England/NORT-28406C/2020\|EPI_ISL_488622\|20 | ···················· | ···················· | ···················· | ···················· | ···················· | ···················· |
| E | 9296 | hCoV-19/England/NORT-283E09/2020\|EPI_ISL_488624\|20 | ···················· | ···················· | ···················· | ···················· | ···················· | ···················· |
| E | 9297 | hCoV-19/England/NORT-295842/2020\|EPI_ISL_488625\|20 | ···················· | ···················· | ···················· | ···················· | ···················· | ···················· |
| E | 9298 | hCoV-19/England/NORT-282110/2020\|EPI_ISL_488626\|20 | ···················· | ···················· | ···················· | ···················· | ···················· | ···················· |
| E | 9299 | hCoV-19/England/NORT-2830F4/2020\|EPI_ISL_488627\|20 | ···················· | ···················· | ···················· | ···················· | ···················· | ···················· |
| E | 9300 | hCoV-19/England/NORT-28422A/2020\|EPI_ISL_488628\|20 | ···················· | ···················· | ···················· | ···················· | ···················· | ···················· |
| E | 9301 | hCoV-19/England/NORT-295B85/2020\|EPI_ISL_488629\|20 | ···················· | ···················· | ···················· | ···················· | ···················· | ···················· |
| E | 9302 | hCoV-19/England/NORT-283B99/2020\|EPI_ISL_488630\|20 | ···················· | ···················· | ···················· | ···················· | ···················· | ···················· |
| E | 9303 | hCoV-19/England/NORT-282EDD/2020\|EPI_ISL_488631\|20 | ···················· | ···················· | ···················· | ···················· | ···················· | ···················· |
| E | 9304 | hCoV-19/England/NORT-283FBB/2020\|EPI_ISL_488632\|20 | ···················· | ···················· | ···················· | ···················· | ···················· | ···················· |
| E | 9305 | hCoV-19/England/NORT-2834DA/2020\|EPI_ISL_488633\|20 | ···················· | ···················· | ···················· | ···················· | ···················· | ···················· |
| E | 9306 | hCoV-19/England/NORT-295BA3/2020\|EPI_ISL_488634\|20 | ···················· | ···················· | ···················· | ···················· | ···················· | ···················· |
| E | 9307 | hCoV-19/England/NORT-2951CF/2020\|EPI_ISL_488635\|20 | ···················· | ···················· | ···················· | ···················· | ···················· | ···················· |
| E | 9308 | hCoV-19/England/NORT-283FD9/2020\|EPI_ISL_488636\|20 | ···················· | ···················· | ···················· | ···················· | ···················· | ···················· |
| E | 9309 | hCoV-19/England/NORT-283DFD/2020\|EPI_ISL_488637\|20 | ···················· | ···················· | ···················· | ···················· | ···················· | ···················· |
| E | 9310 | hCoV-19/England/NORT-294BFF/2020\|EPI_ISL_488638\|20 | ···················· | ·A·················· | ···················· | ···················· | ···················· | ···················· |
| E | 9311 | hCoV-19/England/NORT-282893/2020\|EPI_ISL_488639\|20 | ···················· | ···················· | ···················· | ···················· | ···················· | ···················· |
| E | 9312 | hCoV-19/England/NORT-283AC9/2020\|EPI_ISL_488640\|20 | ···················· | ···················· | ···················· | ···················· | ···················· | ···················· |
| E | 9313 | hCoV-19/England/NORT-295D07/2020\|EPI_ISL_488641\|20 | ···················· | ···················· | ···················· | ···················· | ···················· | ···················· |
| E | 9314 | hCoV-19/England/NORT-295693/2020\|EPI_ISL_488642\|20 | ···················· | ···················· | ···················· | ···················· | ···················· | ···················· |
| E | 9315 | hCoV-19/England/NORT-2839BD/2020\|EPI_ISL_488643\|20 | ···················· | ···················· | ···················· | ···················· | ···················· | ···················· |
| E | 9316 | hCoV-19/England/NORT-294A7A/2020\|EPI_ISL_488644\|20 | ···················· | ···················· | ···················· | ···················· | ···················· | ···················· |
| E | 9317 | hCoV-19/England/NORT-2832D0/2020\|EPI_ISL_488645\|20 | ···················· | ···················· | ···················· | ···················· | ···················· | ···················· |
| E | 9318 | hCoV-19/England/NORT-2828FD/2020\|EPI_ISL_488646\|20 | ···················· | ···················· | ···················· | ···················· | ···················· | ···················· |
| E | 9319 | hCoV-19/England/NORT-282990/2020\|EPI_ISL_488647\|20 | ···················· | ···················· | ···················· | ···················· | ···················· | ···················· |
| E | 9320 | hCoV-19/England/NORT-295C64/2020\|EPI_ISL_488648\|20 | ····G··············· | ···················· | ···················· | ···················· | ···················· | ···················· |
| E | 9321 | hCoV-19/England/NORT-2954B7/2020\|EPI_ISL_488649\|20 | ···················· | ···················· | ···················· | ···················· | ···················· | ···················· |
| E | 9322 | hCoV-19/England/NORT-295A6A/2020\|EPI_ISL_488650\|20 | ···················· | ···················· | ···················· | ···················· | ···················· | ··············A····· |
| E | 9323 | hCoV-19/England/NORT-295684/2020\|EPI_ISL_488651\|20 | ···················· | ···················· | ···················· | ···················· | ···················· | ···················· |
| E | 9324 | hCoV-19/England/NORT-29597C/2020\|EPI_ISL_488652\|20 | ···················· | ·A·················· | ···················· | ···················· | ···················· | ···················· |
| E | 9325 | hCoV-19/England/NORT-28275A/2020\|EPI_ISL_488654\|20 | ···················· | ···················· | ···················· | ···················· | ···················· | ···················· |
| E | 9326 | hCoV-19/England/NORT-28498F/2020\|EPI_ISL_488666\|20 | ···················· | ···················· | ···················· | ···················· | ···················· | ···················· |
| E | 9327 | hCoV-19/England/NORT-2844F7/2020\|EPI_ISL_488667\|20 | ···················· | ···················· | ···················· | ···················· | ···················· | ···················· |
| E | 9328 | hCoV-19/England/NORT-295754/2020\|EPI_ISL_488668\|20 | ···················· | ···················· | ···················· | ···················· | ···················· | ···················· |
| E | 9329 | hCoV-19/England/NORT-2847FE/2020\|EPI_ISL_488669\|20 | ···················· | ···················· | ···················· | ···················· | ···················· | ···················· |
| E | 9330 | hCoV-19/England/NORT-28258D/2020\|EPI_ISL_488670\|20 | ···················· | ···················· | ···················· | ···················· | ···················· | ···················· |
| E | 9331 | hCoV-19/England/NORT-2836D4/2020\|EPI_ISL_488671\|20 | ···················· | ···················· | ···················· | ···················· | ···················· | ···················· |
| E | 9332 | hCoV-19/England/NORT-29548A/2020\|EPI_ISL_488672\|20 | ···················· | ···················· | ···················· | ···················· | ···················· | ···················· |
| E | 9333 | hCoV-19/England/NORT-2956FD/2020\|EPI_ISL_488677\|20 | ···················· | ···················· | ···················· | ···················· | ···················· | ···················· |
| E | 9334 | hCoV-19/England/NORT-283980/2020\|EPI_ISL_488678\|20 | ···················· | ···················· | ···················· | ···················· | ···················· | ···················· |
| E | 9335 | hCoV-19/England/NORT-2949AA/2020\|EPI_ISL_488679\|20 | ···················· | ···················· | ···················· | ···················· | ···················· | ···················· |
| E | 9336 | hCoV-19/England/NORT-282F61/2020\|EPI_ISL_488680\|20 | ···················· | ···················· | ···················· | ···················· | ···················· | ···················· |
| E | 9337 | hCoV-19/England/NORT-2844CA/2020\|EPI_ISL_488681\|20 | ···················· | ···················· | ···················· | ···················· | ···················· | ···················· |
| E | 9338 | hCoV-19/England/NORT-28206E/2020\|EPI_ISL_488682\|20 | ···················· | ···················· | ···················· | ···················· | ···················· | ···················· |
| E | 9339 | hCoV-19/England/NORT-2824BD/2020\|EPI_ISL_488683\|20 | ···················· | ···················· | ···················· | ···················· | ···················· | ···················· |
| E | 9340 | hCoV-19/England/NORT-2838FC/2020\|EPI_ISL_488684\|20 | ···················· | ···················· | ···················· | ···················· | ···················· | ···················· |
| E | 9341 | hCoV-19/England/NORT-295CA0/2020\|EPI_ISL_488685\|20 | ···················· | ···················· | ···················· | ···················· | ···················· | ···················· |
| E | 9342 | hCoV-19/England/NORT-282C79/2020\|EPI_ISL_488686\|20 | ···················· | ···················· | ···················· | ···················· | ···················· | ···················· |
| E | 9343 | hCoV-19/England/NORT-282E55/2020\|EPI_ISL_488687\|20 | ···················· | ···················· | ···················· | ···················· | ···················· | ···················· |
| E | 9344 | hCoV-19/England/NORT-283ABA/2020\|EPI_ISL_488688\|20 | ···················· | ···················· | ···················· | ···················· | ···················· | ···················· |
| E | 9345 | hCoV-19/England/NORT-295095/2020\|EPI_ISL_488689\|20 | ···················· | ···················· | ···················· | ···················· | ···················· | ···················· |
| E | 9346 | hCoV-19/England/NORT-283D2A/2020\|EPI_ISL_488690\|20 | ···················· | ···················· | ···················· | ···················· | ···················· | ···················· |
| E | 9347 | hCoV-19/England/NORT-29595E/2020\|EPI_ISL_488691\|20 | ···················· | ···················· | ···················· | ···················· | ···················· | ···················· |
| E | 9348 | hCoV-19/England/NORT-29588E/2020\|EPI_ISL_488692\|20 | ···················· | ···················· | ···················· | ···················· | ···················· | ···················· |
| E | 9349 | hCoV-19/England/NORT-283926/2020\|EPI_ISL_488693\|20 | ···················· | ···················· | ···················· | ···················· | ···················· | ···················· |
| E | 9350 | hCoV-19/England/NORT-283D1B/2020\|EPI_ISL_488694\|20 | ···················· | ···················· | ···················· | ···················· | ···················· | ···················· |
| E | 9351 | hCoV-19/England/NORT-294F5D/2020\|EPI_ISL_488695\|20 | ···················· | ···················· | ···················· | ···················· | ···················· | ···················· |
| E | 9352 | hCoV-19/England/NORT-29555A/2020\|EPI_ISL_488696\|20 | ···················· | ···················· | ···················· | ···················· | ···················· | ···················· |
| E | 9353 | hCoV-19/England/NORT-295930/2020\|EPI_ISL_488697\|20 | ···················· | ···················· | ···················· | ···················· | ···················· | ···················· |
| E | 9354 | hCoV-19/England/NORT-295B2B/2020\|EPI_ISL_488698\|20 | ···················· | ···················· | ···················· | ···················· | ···················· | ···················· |

| | | | | | | | |
|---|---|---|---|---|---|---|---|
| E | 9355 hCoV-19/USA/MN-GMF-07592/2020\|EPI_ISL_489900\|2020- | ···················· | ···················· | ···················· | ···················· | ···················· | ···················· |
| E | 9356 hCoV-19/USA/MN-GMF-09031/2020\|EPI_ISL_489902\|2020- | ···················· | ···················· | ···················· | ···················· | ···················· | ···················· |
| E | 9357 hCoV-19/USA/MN-GMF-09331/2020\|EPI_ISL_489903\|2020- | ···················· | ·A·················· | ···················· | ···················· | ···················· | ···················· |
| E | 9358 hCoV-19/USA/WI-GMF-06563/2020\|EPI_ISL_489905\|2020- | ···················· | ···················· | ···················· | ···················· | ···················· | ···················· |
| E | 9359 hCoV-19/USA/WI-GMF-06615/2020\|EPI_ISL_489906\|2020- | ···················· | ···················· | ···················· | ···················· | ···················· | ···················· |
| E | 9360 hCoV-19/USA/WI-GMF-06616/2020\|EPI_ISL_489907\|2020- | ···················· | ···················· | ···················· | ···················· | ···················· | ···················· |
| E | 9361 hCoV-19/USA/WI-GMF-06729/2020\|EPI_ISL_489908\|2020- | ···················· | ···················· | ···················· | ···················· | ···················· | ···················· |
| E | 9362 hCoV-19/USA/WI-GMF-06776/2020\|EPI_ISL_489909\|2020- | ···················· | ···················· | ···················· | ···················· | ···················· | ···················· |
| E | 9363 hCoV-19/USA/WI-GMF-06787/2020\|EPI_ISL_489910\|2020- | ···················· | ···················· | ···················· | ···················· | ···················· | ···················· |
| E | 9364 hCoV-19/USA/WI-GMF-06792/2020\|EPI_ISL_489911\|2020- | ···················· | ···················· | ···················· | ···················· | ···················· | ···················· |
| E | 9365 hCoV-19/USA/WI-GMF-06872/2020\|EPI_ISL_489913\|2020- | ···················· | ···················· | ···················· | ···················· | ···················· | ···················· |
| E | 9366 hCoV-19/USA/WI-GMF-06932/2020\|EPI_ISL_489914\|2020- | ···················· | ···················· | ···················· | ···················· | ···················· | ···················· |
| E | 9367 hCoV-19/USA/WI-GMF-07162/2020\|EPI_ISL_489915\|2020- | ···················· | ···················· | ···················· | ···················· | ···················· | ···················· |
| E | 9368 hCoV-19/USA/WI-GMF-07169/2020\|EPI_ISL_489916\|2020- | ············T······· | ···················· | ···················· | ···················· | ···················· | ···················· |
| E | 9369 hCoV-19/USA/WI-GMF-07214/2020\|EPI_ISL_489917\|2020- | ···················· | ···················· | ···················· | ···················· | ···················· | ···················· |
| E | 9370 hCoV-19/USA/WI-GMF-07392/2020\|EPI_ISL_489918\|2020- | ···················· | ···················· | ···················· | ···················· | ···················· | ···················· |
| E | 9371 hCoV-19/USA/WI-GMF-07395/2020\|EPI_ISL_489919\|2020- | ···················· | ···················· | ···················· | ···················· | ···················· | ···················· |
| E | 9372 hCoV-19/USA/WI-GMF-07403/2020\|EPI_ISL_489920\|2020- | ···················· | ···················· | ···················· | ···················· | ···················· | ···················· |
| E | 9373 hCoV-19/USA/WI-GMF-07491/2020\|EPI_ISL_489921\|2020- | ···················· | ···················· | ···················· | ···················· | ···················· | ···················· |
| E | 9374 hCoV-19/USA/WI-GMF-07607/2020\|EPI_ISL_489922\|2020- | ···················· | ···················· | ···················· | ···················· | ···················· | ···················· |
| E | 9375 hCoV-19/USA/WI-GMF-07684/2020\|EPI_ISL_489923\|2020- | ···················· | ···················· | ···················· | ···················· | ···················· | ···················· |
| E | 9376 hCoV-19/USA/WI-GMF-07816/2020\|EPI_ISL_489925\|2020- | ···················· | ···················· | ···················· | ···················· | ···················· | ···················· |
| E | 9377 hCoV-19/USA/WI-GMF-07962/2020\|EPI_ISL_489926\|2020- | ···················· | ···················· | ···················· | ···················· | ···················· | ···················· |
| E | 9378 hCoV-19/USA/WI-GMF-08126/2020\|EPI_ISL_489928\|2020- | ···················· | ···················· | ···················· | ···················· | ···················· | ···················· |
| E | 9379 hCoV-19/USA/WI-GMF-08324/2020\|EPI_ISL_489930\|2020- | ···················· | ···················· | ···················· | ···················· | ···················· | ···················· |
| E | 9380 hCoV-19/USA/WI-GMF-08666/2020\|EPI_ISL_489933\|2020- | ···················· | ···················· | ···················· | ···················· | ···················· | ···················· |
| E | 9381 hCoV-19/USA/WI-GMF-08729/2020\|EPI_ISL_489935\|2020- | ···················· | ···················· | ···················· | ···················· | ···················· | ···················· |
| E | 9382 hCoV-19/USA/WI-GMF-08766/2020\|EPI_ISL_489936\|2020- | ···················· | ···················· | ···················· | ···················· | ···················· | ···················· |
| E | 9383 hCoV-19/USA/WI-GMF-08878/2020\|EPI_ISL_489937\|2020- | ···················· | ···················· | ···················· | ···················· | ···················· | ···················· |
| E | 9384 hCoV-19/USA/WI-GMF-08879/2020\|EPI_ISL_489938\|2020- | ···················· | ···················· | ···················· | ···················· | ···················· | ···················· |
| E | 9385 hCoV-19/USA/WI-GMF-09008/2020\|EPI_ISL_489940\|2020- | ···················· | ···················· | ···················· | ···················· | ···················· | ···················· |
| E | 9386 hCoV-19/USA/WI-GMF-09168/2020\|EPI_ISL_489941\|2020- | ···················· | ···················· | ···················· | ···················· | ···················· | ···················· |
| E | 9387 hCoV-19/USA/WI-GMF-09171/2020\|EPI_ISL_489942\|2020- | ···················· | ···················· | ···················· | ···················· | ···················· | ···················· |
| E | 9388 hCoV-19/USA/WI-GMF-09205/2020\|EPI_ISL_489943\|2020- | ···················· | ···················· | ···················· | ···················· | ···················· | ···················· |
| E | 9389 hCoV-19/USA/WI-GMF-09471/2020\|EPI_ISL_489945\|2020- | ···················· | ···················· | ···················· | ···················· | ···················· | ···················· |
| E | 9390 hCoV-19/USA/WI-GMF-09489/2020\|EPI_ISL_489946\|2020- | ···················· | ·A·················· | ···················· | ···················· | ···················· | ···················· |
| E | 9391 hCoV-19/USA/WI-GMF-09509/2020\|EPI_ISL_489948\|2020- | ···················· | ···················· | ···················· | ···················· | ···················· | ···················· |
| E | 9392 hCoV-19/USA/WI-GMF-09554/2020\|EPI_ISL_489949\|2020- | ···················· | ···················· | ···················· | ···················· | ···················· | ···················· |
| E | 9393 hCoV-19/USA/WI-GMF-09661/2020\|EPI_ISL_489950\|2020- | ···················· | ···················· | ···················· | ···················· | ···················· | ···················· |
| E | 9394 hCoV-19/USA/WI-GMF-09664/2020\|EPI_ISL_489951\|2020- | ···················· | ···················· | ···················· | ···················· | ···················· | ···················· |
| E | 9395 hCoV-19/USA/WI-GMF-09680/2020\|EPI_ISL_489952\|2020- | ···················· | ···················· | ···················· | ···················· | ···················· | ···················· |
| E | 9396 hCoV-19/USA/WI-GMF-09683/2020\|EPI_ISL_489953\|2020- | ···················· | ·A·················· | ···················· | ···················· | ···················· | ···················· |
| E | 9397 hCoV-19/USA/WI-GMF-09715/2020\|EPI_ISL_489954\|2020- | ···················· | ···················· | ···················· | ···················· | ···················· | ···················· |
| E | 9398 hCoV-19/USA/WI-GMF-09752/2020\|EPI_ISL_489955\|2020- | ···················· | ···················· | ···················· | ···················· | ···················· | ···················· |
| E | 9399 hCoV-19/USA/WI-GMF-M00008/2020\|EPI_ISL_489956\|2020 | ···················· | ···················· | ···················· | ···················· | ···················· | ···················· |
| E | 9400 hCoV-19/USA/WI-GMF-M00011/2020\|EPI_ISL_489957\|2020 | ···················· | ···················· | ···················· | ···················· | ···················· | ···················· |
| E | 9401 hCoV-19/USA/WI-GMF-M00012/2020\|EPI_ISL_489958\|2020 | ···················· | ···················· | ···················· | ···················· | ···················· | ···················· |
| E | 9402 hCoV-19/Switzerland/AG-ETHZ-180003/2020\|EPI_ISL_48 | ···················· | ···················· | ···················· | ···················· | ···················· | ···················· |
| E | 9403 hCoV-19/Switzerland/BE-ETHZ-180004/2020\|EPI_ISL_48 | ···················· | ···················· | ···················· | ···················· | ···················· | ···················· |
| E | 9404 hCoV-19/Switzerland/GR-ETHZ-180005/2020\|EPI_ISL_48 | ···················· | ···················· | ···················· | ···················· | ···················· | ···················· |
| E | 9405 hCoV-19/Switzerland/ZH-ETHZ-180007/2020\|EPI_ISL_48 | ···················· | ···················· | ···················· | ···················· | ···················· | ···················· |
| E | 9406 hCoV-19/Switzerland/FR-ETHZ-180008/2020\|EPI_ISL_48 | ···················· | ···················· | ···················· | ···················· | ···················· | ···················· |
| E | 9407 hCoV-19/Switzerland/VS-ETHZ-180011/2020\|EPI_ISL_48 | ···················· | ···················· | ···················· | ···················· | ···················· | ···················· |
| E | 9408 hCoV-19/Switzerland/AG-ETHZ-180018/2020\|EPI_ISL_48 | ···················· | ···················· | ···················· | ···················· | ···················· | ···················· |
| E | 9409 hCoV-19/Switzerland/BE-ETHZ-180019/2020\|EPI_ISL_48 | ···················· | ···················· | ···················· | ···················· | ···················· | ···················· |
| E | 9410 hCoV-19/Switzerland/SG-ETHZ-180020/2020\|EPI_ISL_48 | ···················· | ···················· | ···················· | ···················· | ···················· | ···················· |
| E | 9411 hCoV-19/Switzerland/SZ-ETHZ-180021/2020\|EPI_ISL_48 | ···················· | ···················· | ···················· | ···················· | ···················· | ···················· |
| E | 9412 hCoV-19/Switzerland/AG-ETHZ-180023/2020\|EPI_ISL_48 | ···················· | ···················· | ···················· | ···················· | ···················· | ···················· |
| E | 9413 hCoV-19/Switzerland/ZH-ETHZ-180024/2020\|EPI_ISL_48 | ···················· | ···················· | ···················· | ···················· | ···················· | ···················· |
| E | 9414 hCoV-19/Switzerland/SG-ETHZ-180030/2020\|EPI_ISL_48 | ···················· | ···················· | ···················· | ···················· | ···················· | ···················· |
| E | 9415 hCoV-19/Switzerland/ZH-ETHZ-180031/2020\|EPI_ISL_48 | ···················· | ···················· | ···················· | ···················· | ···················· | ···················· |
| E | 9416 hCoV-19/Switzerland/SH-ETHZ-180032/2020\|EPI_ISL_48 | ···················· | ···················· | ···················· | ···················· | ···················· | ···················· |
| E | 9417 hCoV-19/Switzerland/ZH-ETHZ-180037/2020\|EPI_ISL_48 | ···················· | ···················· | ···················· | ···················· | ···················· | ···················· |
| E | 9418 hCoV-19/Switzerland/BL-ETHZ-180038/2020\|EPI_ISL_48 | ···················· | ···················· | ···················· | ···················· | ···················· | ···················· |
| E | 9419 hCoV-19/Switzerland/VD-ETHZ-180039/2020\|EPI_ISL_48 | ···················· | ···················· | ···················· | ···················· | ···················· | ···················· |
| E | 9420 hCoV-19/Switzerland/ZH-ETHZ-180040/2020\|EPI_ISL_48 | ···················· | ···················· | ···················· | ···················· | ···················· | ···················· |
| E | 9421 hCoV-19/Switzerland/ZH-ETHZ-180041/2020\|EPI_ISL_48 | ···················· | ···················· | ···················· | ···················· | ···················· | ··········R········· |
| E | 9422 hCoV-19/Switzerland/ZH-ETHZ-180042/2020\|EPI_ISL_48 | ···················· | ···················· | ···················· | ···················· | ···················· | ···················· |
| E | 9423 hCoV-19/Switzerland/ZH-ETHZ-180043/2020\|EPI_ISL_48 | ···················· | ···················· | ···················· | ···················· | ···················· | ···················· |
| E | 9424 hCoV-19/Switzerland/SZ-ETHZ-180045/2020\|EPI_ISL_48 | ···················· | ···················· | ···················· | ···················· | ···················· | ···················· |
| E | 9425 hCoV-19/Switzerland/ZH-ETHZ-180047/2020\|EPI_ISL_48 | ···················· | ···················· | ···················· | ···················· | ···················· | ···················· |
| E | 9426 hCoV-19/Switzerland/UR-ETHZ-180049/2020\|EPI_ISL_48 | ···········T········ | ···················· | ···················· | ···················· | ···················· | ···················· |
| E | 9427 hCoV-19/Switzerland/UR-ETHZ-180051/2020\|EPI_ISL_48 | ···········T········ | ···················· | ···················· | ···················· | ···················· | ···················· |
| E | 9428 hCoV-19/Saudi Arabia/492/2020\|EPI_ISL_489997\|2020- | ···················· | ···················· | ···················· | ···················· | ···················· | ···················· |
| E | 9429 hCoV-19/Saudi Arabia/509/2020\|EPI_ISL_489998\|2020- | ···················· | ···················· | ···················· | ···················· | ···················· | ···················· |
| E | 9430 hCoV-19/Saudi Arabia/518/2020\|EPI_ISL_489999\|2020- | ···················· | ···················· | ···················· | ···················· | ···················· | ···················· |
| E | 9431 hCoV-19/Spain/PV-HUB-56574576/2020\|EPI_ISL_490977\| | ···················· | ···················· | ···················· | ···················· | ···················· | ···················· |
| E | 9432 hCoV-19/USA/AR-UW-MAYO83/2020\|EPI_ISL_490978\|2020- | ···················· | ···················· | ···················· | ···················· | ···················· | ···················· |
| E | 9433 hCoV-19/USA/CT-UW-3109/2020\|EPI_ISL_490979\|2020-03 | ···················· | ···················· | ···················· | ···················· | ···················· | ···················· |
| E | 9434 hCoV-19/USA/ID-UW-3183/2020\|EPI_ISL_490990\|2020-03 | ···················· | ···················· | ···················· | ···················· | ···················· | ···················· |

E 9435 hCoV-19/USA/ID-UW-3184/2020|EPI_ISL_490991|2020-03 ···················· ···················· ····················· ···················· ···················· ····················
E 9436 hCoV-19/USA/ID-UW-3196/2020|EPI_ISL_490992|2020-03 ···················· ···················· ····················· ···················· ···················· ····················
E 9437 hCoV-19/USA/ID-UW-3204/2020|EPI_ISL_490993|2020-03 ···················· ···················· ····················· ···················· ···················· ····················
E 9438 hCoV-19/USA/ID-UW-3335/2020|EPI_ISL_490994|2020-03 ···················· ···················· ····················· ···················· ···················· ····················
E 9439 hCoV-19/USA/ID-UW-3416/2020|EPI_ISL_490995|2020-03 ···················· ···················· ····················· ···················· ···················· ····················
E 9440 hCoV-19/USA/ID-UW-3481/2020|EPI_ISL_490996|2020-03 ···················· ···················· ····················· ···················· ···················· ····················
E 9441 hCoV-19/USA/LA-UW-MAYO81/2020|EPI_ISL_490997|2020- ···················· ···················· ····················· ···················· ···················· ····················
E 9442 hCoV-19/USA/LA-UW-MAYO82/2020|EPI_ISL_490998|2020- ···················· ···················· ····················· ···················· ···················· ····················
E 9443 hCoV-19/USA/MA-UW-MAYO44/2020|EPI_ISL_490999|2020- ···················· ···················· ····················· ···················· ···················· ····················
E 9444 hCoV-19/Wuhan/0125-A191/2020|EPI_ISL_493165|2020-0 ···················· ···················· ····················· ···················· ···················· ····················
E 9445 hCoV-19/Wuhan/0125-A192/2020|EPI_ISL_493166|2020-0 ···················· ···················· ····················· ···················· ···················· ····················
E 9446 hCoV-19/Wuhan/0126-C5/2020|EPI_ISL_493167|2020-01- ···················· ···················· ····················· ···················· ···················· ····················
E 9447 hCoV-19/Wuhan/0126-C7/2020|EPI_ISL_493168|2020-01- ···················· ···················· ····················· ···················· ···················· ····················
E 9448 hCoV-19/Wuhan/0126-C10/2020|EPI_ISL_493169|2020-01 ···················· ···················· ····················· ···················· ···················· ····················
E 9449 hCoV-19/Wuhan/0126-C12/2020|EPI_ISL_493170|2020-01 ···················· ···················· ····················· ···················· ···················· ····················
E 9450 hCoV-19/Wuhan/0126-C13/2020|EPI_ISL_493171|2020-01 ···················· ···················· ····················· ···················· ···················· ····················
E 9451 hCoV-19/Wuhan/0126-C14/2020|EPI_ISL_493172|2020-01 ···················· ···················· ····················· ···················· ···················· ····················
E 9452 hCoV-19/Wuhan/0126-C26/2020|EPI_ISL_493173|2020-01 ···················· ···················· ····················· ···················· ···················· ····················
E 9453 hCoV-19/Wuhan/0126-C31/2020|EPI_ISL_493174|2020-01 ···················· ···················· ····················· ···················· ···················· ····················
E 9454 hCoV-19/Wuhan/0126-C43/2020|EPI_ISL_493175|2020-01 ···················· ···················· ····················· ···················· ···················· ····················
E 9455 hCoV-19/Wuhan/0126-C45/2020|EPI_ISL_493176|2020-01 ···················· ···················· ····················· ···················· ···················· ····················
E 9456 hCoV-19/Wuhan/0126-C47/2020|EPI_ISL_493177|2020-01 ···················· ···················· ····················· ···················· ···················· ····················
E 9457 hCoV-19/Wuhan/0126-C77/2020|EPI_ISL_493178|2020-01 ···················· ···················· ····················· ···················· ···················· ····················
E 9458 hCoV-19/Wuhan/0126-C93/2020|EPI_ISL_493179|2020-01 ···················· ···················· ····················· ···················· ···················· ····················
E 9459 hCoV-19/Wuhan/0126-C94/2020|EPI_ISL_493180|2020-01 ···················· ···················· ····················· ···················· ···················· ····················
E 9460 hCoV-19/Wuhan/0126-C98/2020|EPI_ISL_493181|2020-01 ···················· ···················· ····················· ···················· ···················· ····················
E 9461 hCoV-19/Wuhan/0126-C100/2020|EPI_ISL_493182|2020-0 ···················· ···················· ····················· ···················· ···················· ····················
E 9462 hCoV-19/Wuhan/0126-C101/2020|EPI_ISL_493183|2020-0 ···················· ···················· ····················· ···················· ···················· ····················
E 9463 hCoV-19/Wuhan/0126-C102/2020|EPI_ISL_493184|2020-0 ···················· ···················· ····················· ···················· ···················· ····················
E 9464 hCoV-19/Wuhan/0126-C103/2020|EPI_ISL_493185|2020-0 ···················· ···················· ····················· ···················· ···················· ····················
E 9465 hCoV-19/Wuhan/0126-C106/2020|EPI_ISL_493186|2020-0 ···················· ···················· ····················· ···················· ···················· ····················
E 9466 hCoV-19/Wuhan/0126-C109/2020|EPI_ISL_493188|2020-0 ···················· ···················· ····················· ···················· ···················· ····················
E 9467 hCoV-19/Wuhan/0126-C111/2020|EPI_ISL_493189|2020-0 ···················· ···················· ····················· ···················· ···················· ····················
E 9468 hCoV-19/Taiwan/NCKUH-002/2020|EPI_ISL_493198|2020- ···················· ···················· ····················· ···················· ···················· ····················
E 9469 hCoV-19/Taiwan/NCKUH-003/2020|EPI_ISL_493199|2020- ···················· ···················· ····················· ···················· ···················· ····················
E 9470 hCoV-19/USA/CA-ALSR-1332/2020|EPI_ISL_494496|2020- ···················· ···················· ····················· ···················· ···················· ····················
E 9471 hCoV-19/USA/CA-ALSR-1333/2020|EPI_ISL_494497|2020- ···················· ···················· ····················· ···················· ···················· ····················
E 9472 hCoV-19/USA/CA-ALSR-1335/2020|EPI_ISL_494499|2020- ···················· ···················· ····················· ···················· ···················· ····················
E 9483 hCoV-19/India/OR-ILSCV25367/2020|EPI_ISL_481110|20 ···················· ···················· ····················· ···················· ···················· ····················
E 9484 hCoV-19/India/OR-ILSCV25378/2020|EPI_ISL_481111|20 ···················· ···················· ····················· ···················· ···················· ····················
E 9485 hCoV-19/India/OR-ILSCV25518/2020|EPI_ISL_481112|20 ···················· ···················· ····················· ···················· ···················· ····················
E 9486 hCoV-19/India/OR-ILSCV25524/2020|EPI_ISL_481113|20 ···················· ···················· ····················· ···················· ···················· ····················
E 9487 hCoV-19/India/OR-ILSCV25591/2020|EPI_ISL_481114|20 ···················· ···················· ····················· ···················· ···················· ····················
E 9488 hCoV-19/India/OR-ILSCV25594/2020|EPI_ISL_481115|20 ···················· ···················· ····················· ···················· ···················· ····················
E 9489 hCoV-19/India/OR-ILSCV26793/2020|EPI_ISL_481116|20 ···················· ···················· ····················· ···················· ···················· ····················
E 9490 hCoV-19/India/OR-ILSCV26795/2020|EPI_ISL_481117|20 ···················· ···················· ····················· ···················· ···················· ····················
E 9491 hCoV-19/India/OR-ILSCV27381/2020|EPI_ISL_481118|20 ···················· ···················· ····················· ···················· ···················· ····················
E 9492 hCoV-19/India/OR-ILSCV27406/2020|EPI_ISL_481119|20 ···················· ···················· ····················· ···················· ···················· ····················
E 9493 hCoV-19/India/OR-ILSCV27409/2020|EPI_ISL_481120|20 ···················· ···················· ····················· ···················· ···················· ····················
E 9494 hCoV-19/India/OR-ILSCV27713/2020|EPI_ISL_481121|20 ···················· ···················· ····················· ···················· ···················· ····················
E 9495 hCoV-19/India/OR-ILSCV27725/2020|EPI_ISL_481122|20 ···················· ···················· ····················· ···················· ···················· ····················
E 9496 hCoV-19/India/OR-ILSCV27726/2020|EPI_ISL_481123|20 ···················· ···················· ····················· ···················· ···················· ····················
E 9497 hCoV-19/India/OR-ILSCV29666/2020|EPI_ISL_481130|20 ···················· ···················· ····················· ···················· ···················· ····················
E 9498 hCoV-19/India/OR-ILSCV30167/2020|EPI_ISL_481142|20 ···················· ···················· ····················· ···················· ···················· ····················
E 9499 hCoV-19/India/OR-ILSCV30220/2020|EPI_ISL_481143|20 ···················· ···················· ····················· ···················· ···················· ····················
E 9500 hCoV-19/India/OR-ILSCV30221/2020|EPI_ISL_481144|20 ···················· ···················· ····················· ···················· ···················· ····················
E 9501 hCoV-19/India/OR-ILSCV30442/2020|EPI_ISL_481146|20 ···················· ···················· ····················· ···················· ···················· ····················
E 9502 hCoV-19/India/OR-ILSCV31185/2020|EPI_ISL_481147|20 ···················· ···················· ····················· ···················· ···················· ····················
E 9503 hCoV-19/India/OR-ILSCV31447/2020|EPI_ISL_481148|20 ···················· ·A·················· ····················· ···················· ···················· ··A·················
E 9504 hCoV-19/India/OR-ILSCV31650/2020|EPI_ISL_481149|20 ···················· ···················· ····················· ···················· ···················· ····················
E 9505 hCoV-19/India/OR-ILSCV31684/2020|EPI_ISL_481150|20 ···················· ···················· ····················· ···················· ···················· ····················
E 9506 hCoV-19/India/OR-ILSCV31693/2020|EPI_ISL_481151|20 ···················· ···················· ····················· ···················· ···················· ····················
E 9507 hCoV-19/India/OR-ILSCV31694/2020|EPI_ISL_481152|20 ···················· ···················· ····················· ···················· ···················· ····················
E 9508 hCoV-19/India/OR-ILSCV31984/2020|EPI_ISL_481153|20 ···················· ···················· ····················· ···················· ···················· ····················
E 9509 hCoV-19/India/OR-ILSCV32007/2020|EPI_ISL_481154|20 ···················· ···················· ····················· ···················· ···················· ····················
E 9510 hCoV-19/India/OR-ILSCV32142/2020|EPI_ISL_481155|20 ···················· ···················· ····················· ···················· ···················· ····················
E 9511 hCoV-19/India/OR-ILSCV32465/2020|EPI_ISL_481156|20 ···················· ···················· ····················· ···················· ···················· ····················
E 9512 hCoV-19/India/OR-ILSCV32485/2020|EPI_ISL_481157|20 ···················· ···················· ····················· ···················· ···················· ····················
E 9513 hCoV-19/India/OR-ILSCV32486/2020|EPI_ISL_481158|20 ···················· ···················· ····················· ···················· ···················· ····················
E 9514 hCoV-19/India/OR-ILSCV32593/2020|EPI_ISL_481159|20 ···················· ···················· ····················· ···················· ···················· ····················
E 9515 hCoV-19/India/OR-ILSCV32652/2020|EPI_ISL_481160|20 ···················· ·A·················· ····················· ···················· ···················· ····················
E 9516 hCoV-19/India/OR-ILSCV32656/2020|EPI_ISL_481161|20 ···················· ···················· ····················· ···················· ···················· ····················
E 9517 hCoV-19/India/OR-ILSCV32670/2020|EPI_ISL_481162|20 ···················· ···················· ····················· ···················· ···················· ····················
E 9518 hCoV-19/India/OR-ILSCV32814/2020|EPI_ISL_481163|20 ···················· ···················· ····················· ···················· ···················· ····················
E 9519 hCoV-19/India/OR-ILSCV32815/2020|EPI_ISL_481164|20 ···················· ···················· ····················· ···················· ···················· ····················
E 9520 hCoV-19/India/OR-ILSCV32816/2020|EPI_ISL_481165|20 ···················· ···················· ····················· ···················· ···················· ····················
E 9521 hCoV-19/India/OR-ILSCV32817/2020|EPI_ISL_481166|20 ···················· ···················· ····················· ···················· ···················· ····················
E 9522 hCoV-19/India/OR-ILSCV32866/2020|EPI_ISL_481167|20 ···················· ···················· ····················· ···················· ···················· ····················
E 9523 hCoV-19/India/OR-ILSCV32870/2020|EPI_ISL_481168|20 ···················· ···················· ····················· ···················· ···················· ····················
E 9524 hCoV-19/India/OR-ILSCV32872/2020|EPI_ISL_481169|20 ···················· ···················· ····················· ···················· ···················· ····················

```
E  9525 hCoV-19/India/OR-ILSCV32879/2020|EPI_ISL_481170|20  ····················  ····················  ····················  ····················  ····················  ····················
E  9526 hCoV-19/India/OR-ILSCV32889/2020|EPI_ISL_481171|20  ····················  ····················  ····················  ····················  ····················  ····················
E  9527 hCoV-19/India/OR-ILSCV32892/2020|EPI_ISL_481172|20  ····················  ····················  ····················  ····················  ····················  ····················
E  9528 hCoV-19/India/OR-ILSCV32893/2020|EPI_ISL_481173|20  ····················  ····················  ····················  ····················  ····················  ····················
E  9529 hCoV-19/India/OR-ILSCV32895/2020|EPI_ISL_481174|20  ····················  ····················  ····················  ····················  ····················  ····················
E  9530 hCoV-19/India/OR-ILSCV32896/2020|EPI_ISL_481175|20  ····················  ····················  ····················  ····················  ····················  ····················
E  9531 hCoV-19/India/OR-ILSCV32898/2020|EPI_ISL_481176|20  ····················  ····················  ····················  ····················  ····················  ····················
E  9532 hCoV-19/India/OR-ILSCV32899/2020|EPI_ISL_481177|20  ····················  ····················  ····················  ····················  ····················  ····················
E  9533 hCoV-19/India/OR-ILSCV33971/2020|EPI_ISL_481178|20  ····················  ····················  ····················  ····················  ····················  ····················
E  9534 hCoV-19/India/OR-ILSCV33972/2020|EPI_ISL_481179|20  ····················  ····················  ····················  ····················  ····················  ····················
E  9535 hCoV-19/India/OR-ILSCV33982/2020|EPI_ISL_481180|20  ····················  ····················  ····················  ····················  ····················  ····················
E  9536 hCoV-19/India/OR-ILSCV33983/2020|EPI_ISL_481181|20  ····················  ····················  ····················  ····················  ······T·············  ····················
E  9537 hCoV-19/India/OR-ILSCV34478/2020|EPI_ISL_481182|20  ····················  ····················  ····················  ····················  ····················  ····················
E  9538 hCoV-19/India/OR-ILSCV34479/2020|EPI_ISL_481183|20  ····················  ····················  ····················  ····················  ······T·············  ····················
E  9539 hCoV-19/India/OR-ILSCV34480/2020|EPI_ISL_481184|20  ····················  ····················  ····················  ····················  ····················  ····················
E  9540 hCoV-19/India/OR-ILSCV34933/2020|EPI_ISL_481185|20  ····················  ····················  ····················  ····················  ····················  ····················
E  9541 hCoV-19/USA/OR-PROV-182/2020|EPI_ISL_482430|2020-0  ····················  ····················  ····················  ····················  ····················  ····················
E  9542 hCoV-19/USA/OR-PROV-183/2020|EPI_ISL_482431|2020-0  ····················  ····················  ····················  ····················  ····················  ····················
E  9543 hCoV-19/USA/OR-PROV-184/2020|EPI_ISL_482432|2020-0  ····················  ····················  ····················  ····················  ····················  ····················
E  9544 hCoV-19/USA/OR-PROV-185/2020|EPI_ISL_482433|2020-0  ····················  ····················  ····················  ····················  ····················  ····················
E  9545 hCoV-19/USA/OR-PROV-186/2020|EPI_ISL_482434|2020-0  ····················  ····················  ····················  ····················  ····················  ····················
E  9546 hCoV-19/USA/OR-PROV-187/2020|EPI_ISL_482435|2020-0  ····················  ····················  ····················  ····················  ····················  ····················
E  9547 hCoV-19/USA/OR-PROV-188/2020|EPI_ISL_482436|2020-0  ····················  ····················  ····················  ····················  ····················  ····················
E  9548 hCoV-19/USA/OR-PROV-192/2020|EPI_ISL_482437|2020-0  ····················  ····················  ····················  ····················  ····················  ····················
E  9549 hCoV-19/USA/OR-PROV-193/2020|EPI_ISL_482438|2020-0  ····················  ····················  ····················  ····················  ····················  ····················
E  9550 hCoV-19/USA/OR-PROV-194/2020|EPI_ISL_482439|2020-0  ····················  ····················  ····················  ····················  ····················  ····················
E  9551 hCoV-19/USA/OR-PROV-197/2020|EPI_ISL_482440|2020-0  ····················  ····················  ····················  ····················  ····················  ····················
E  9552 hCoV-19/USA/OR-PROV-199/2020|EPI_ISL_482441|2020-0  ····················  ····················  ····················  ····················  ····················  ····················
E  9553 hCoV-19/USA/OR-PROV-200/2020|EPI_ISL_482442|2020-0  ····················  ····················  ····················  ····················  ····················  ····················
E  9554 hCoV-19/USA/OR-PROV-201/2020|EPI_ISL_482443|2020-0  ····················  ····················  ····················  ····················  ····················  ····················
E  9555 hCoV-19/USA/OR-PROV-202/2020|EPI_ISL_482444|2020-0  ····················  ····················  ····················  ····················  ····················  ····················
E  9556 hCoV-19/USA/OR-PROV-203/2020|EPI_ISL_482445|2020-0  ····················  ····················  ····················  ····················  ····················  ····················
E  9557 hCoV-19/USA/OR-PROV-204/2020|EPI_ISL_482446|2020-0  ····················  ····················  ····················  ····················  ····················  ····················
E  9558 hCoV-19/USA/OR-PROV-205/2020|EPI_ISL_482447|2020-0  ····················  ····················  ····················  ····················  ····················  ····················
E  9559 hCoV-19/USA/OR-PROV-206/2020|EPI_ISL_482448|2020-0  ····················  ····················  ····················  ····················  ····················  ····················
E  9560 hCoV-19/USA/OR-PROV-207/2020|EPI_ISL_482449|2020-0  ····················  ····················  ····················  ····················  ····················  ····················
E  9561 hCoV-19/USA/OR-PROV-209/2020|EPI_ISL_482450|2020-0  ····················  ····················  ····················  ····················  ····················  ····················
E  9562 hCoV-19/USA/OR-PROV-216/2020|EPI_ISL_482451|2020-0  ····················  ····················  ····················  ····················  ····················  ····················
E  9563 hCoV-19/USA/OR-PROV-217/2020|EPI_ISL_482452|2020-0  ····················  ····················  ····················  ····················  ····················  ····················
E  9564 hCoV-19/USA/OR-PROV-218/2020|EPI_ISL_482453|2020-0  ····················  ····················  ····················  ····················  ····················  ····················
E  9565 hCoV-19/USA/OR-PROV-219/2020|EPI_ISL_482454|2020-0  ····················  ····················  ····················  ····················  ····················  ····················
E  9566 hCoV-19/USA/OR-PROV-225/2020|EPI_ISL_482460|2020-0  ····················  ····················  ····················  ····················  ····················  ····················
E  9567 hCoV-19/USA/OR-PROV-226/2020|EPI_ISL_482461|2020-0  ····················  ····················  ····················  ····················  ····················  ····················
E  9568 hCoV-19/Canada/NB_56/2020|EPI_ISL_482473|2020-03-1  ····················  ····················  ····················  ····················  ····················  ····················
E  9569 hCoV-19/Canada/MB_58/2020|EPI_ISL_482474|2020-03-1  ····················  ····················  ····················  ····················  ····················  ····················
E  9570 hCoV-19/Canada/MB_59/2020|EPI_ISL_482475|2020-03-1  ····················  ····················  ····················  ····················  ····················  ····················
E  9571 hCoV-19/Canada/NS_60/2020|EPI_ISL_482476|2020-03-1  ····················  ····················  ····················  ····················  ····················  ····················
E  9572 hCoV-19/Canada/NS_61/2020|EPI_ISL_482477|2020-03-1  ····················  ····················  ····················  ····················  ····················  ····················
E  9573 hCoV-19/Canada/MB_71/2020|EPI_ISL_482478|2020-03-1  ····················  ····A···············  ····················  ····················  ····················  ····················
E  9574 hCoV-19/Canada/NL_72/2020|EPI_ISL_482479|2020-03-1  ····················  ····················  ····················  ····················  ····················  ····················
E  9575 hCoV-19/Canada/MB_75/2020|EPI_ISL_482480|2020-03-2  ····················  ····················  ····················  ····················  ····················  ····················
E  9576 hCoV-19/Canada/MB_76/2020|EPI_ISL_482481|2020-03-1  ····················  ····················  ····················  ····················  ····················  ····················
E  9577 hCoV-19/Canada/MB_78/2020|EPI_ISL_482482|2020-03-1  ····················  ····················  ····················  ····················  ····················  ····················
E  9578 hCoV-19/Canada/MB_79/2020|EPI_ISL_482483|2020-03-1  ····················  ····················  ····················  ····················  ····················  ····················
E  9579 hCoV-19/Canada/MB_80/2020|EPI_ISL_482484|2020-03-1  ····················  ····················  ····················  ····················  ····················  ····················
E  9580 hCoV-19/Bangladesh/BCSIR-NILMRC-155/2020|EPI_ISL_4  ····················  ····················  ····················  ····················  ····················  ····················
E  9581 hCoV-19/Bangladesh/BCSIR-NILMRC-160/2020|EPI_ISL_4  ····················  ····················  ····················  ····················  ····················  ····················
E  9582 hCoV-19/Bangladesh/BCSIR-NILMRC-118a/2020|EPI_ISL_  ····················  ····················  ····················  ····················  ····················  ····················
E  9583 hCoV-19/Bangladesh/BCSIR-NILMRC-161/2020|EPI_ISL_4  ····················  ····················  ····················  ····················  ····················  ····················
E  9584 hCoV-19/Bangladesh/BCSIR-NILMRC-149/2020|EPI_ISL_4  ····················  ····················  ····················  ····················  ····················  ····················
E  9585 hCoV-19/India/DL-NCDC1580-CSIR-IGIB/2020|EPI_ISL_4  ····················  ·A··················  ····················  ····················  ····················  ···R················
E  9586 hCoV-19/England/NORT-284891/2020|EPI_ISL_488600|20  ····················  ····················  ····················  ····················  ····················  ····················
E  9587 hCoV-19/England/NORT-284A40/2020|EPI_ISL_488602|20  ····················  ····················  ····················  ····················  ····················  ····················
E  9588 hCoV-19/England/NORT-282A9D/2020|EPI_ISL_488603|20  ····················  ····················  ····················  ····················  ····················  ····················
E  9589 hCoV-19/England/NORT-284196/2020|EPI_ISL_488608|20  ····················  ····················  ····················  ····················  ····················  ····················
E  9590 hCoV-19/England/NORT-28256F/2020|EPI_ISL_488609|20  ····················  ····················  ····················  ····················  ····················  ····················
E  9591 hCoV-19/USA/WI-GMF-08470/2020|EPI_ISL_489931|2020-  ····················  ····················  ····················  ····················  ····················  ····················
E  9592 hCoV-19/USA/WI-GMF-08638/2020|EPI_ISL_489932|2020-  ····················  ····················  ····················  ····················  ····················  ····················
E  9593 hCoV-19/USA/WI-GMF-08700/2020|EPI_ISL_489934|2020-  ····················  ····················  ····················  ····················  ····················  ····················
E  9594 hCoV-19/USA/WI-GMF-08889/2020|EPI_ISL_489939|2020-  ····················  ····················  ····················  ····················  ····················  ····················
E  9595 hCoV-19/USA/UT-02528/2020|EPI_ISL_493113|2020-04-2  ····················  ····················  ····················  ····················  ····················  ····················
E  9596 hCoV-19/USA/UT-02530/2020|EPI_ISL_493114|2020-04-2  ····················  ····················  ····················  ····················  ····················  ····················
E  9597 hCoV-19/USA/UT-02541/2020|EPI_ISL_493116|2020-04-2  ····················  ····················  ····················  ····················  ····················  ····················
E  9598 hCoV-19/USA/UT-06242/2020|EPI_ISL_493123|2020-04-2  ····················  ····················  ····················  ····················  ····················  ····················
E  9599 hCoV-19/USA/WY-WYPHL-20007411/2020|EPI_ISL_493125|  ····················  ····················  ····················  ····················  ····················  ····················
E  9600 hCoV-19/USA/WY-WYPHL-20007413/2020|EPI_ISL_493126|  ····················  ····················  ····················  ····················  ····················  ····················
E  9601 hCoV-19/USA/WY-WYPHL-20007414/2020|EPI_ISL_493127|  ····················  ····················  ····················  ····················  ····················  ····················
E  9602 hCoV-19/USA/WY-WYPHL-20007680/2020|EPI_ISL_493128|  ····················  ····················  ····················  ····················  ····················  ····················
E  9603 hCoV-19/USA/SC-USC-COM0003/2020|EPI_ISL_493129|202  ····················  ·········A··········  ····················  ····················  ····················  ····················
E  9604 hCoV-19/Thailand/MUMT-3/2020|EPI_ISL_493137|2020-0  ····················  ····················  ····················  ····················  ····················  ····················
```

| | | | | | | | |
|---|---|---|---|---|---|---|---|
| E | 9605 hCoV-19/Thailand/MUMT-4/2020\|EPI_ISL_493139\|2020-0 | ···················· | ···················· | ····················· | ···················· | ···················· | ···················· |
| E | 9606 hCoV-19/Wuhan/0125-A137/2020\|EPI_ISL_493149\|2020-0 | ···················· | ···················· | ····················· | ···················· | ···················· | ···················· |
| E | 9607 hCoV-19/Wuhan/0125-A142/2020\|EPI_ISL_493150\|2020-0 | ···················· | ···················· | ····················· | ···················· | ···················· | ···················· |
| E | 9608 hCoV-19/Wuhan/0125-A145/2020\|EPI_ISL_493151\|2020-0 | ···················· | ···················· | ····················· | ···················· | ···················· | ···················· |
| E | 9609 hCoV-19/Wuhan/0125-A148/2020\|EPI_ISL_493152\|2020-0 | ···················· | ···················· | ····················· | ···················· | ···················· | ···················· |
| E | 9610 hCoV-19/Wuhan/0125-A152/2020\|EPI_ISL_493153\|2020-0 | ···················· | ···················· | ····················· | ···················· | ···················· | ···················· |
| E | 9611 hCoV-19/Wuhan/0125-A153/2020\|EPI_ISL_493154\|2020-0 | ···················· | ···················· | ····················· | ···················· | ···················· | ···················· |
| E | 9612 hCoV-19/Wuhan/0125-A160/2020\|EPI_ISL_493155\|2020-0 | ···················· | ···················· | ····················· | ···················· | ···················· | ···················· |
| E | 9613 hCoV-19/Wuhan/0125-A163/2020\|EPI_ISL_493156\|2020-0 | ···················· | ···················· | ····················· | ···················· | ···················· | ···················· |
| E | 9614 hCoV-19/Wuhan/0125-A164/2020\|EPI_ISL_493157\|2020-0 | ···················· | ···················· | ····················· | ···················· | ···················· | ···················· |
| E | 9615 hCoV-19/Wuhan/0125-A167/2020\|EPI_ISL_493158\|2020-0 | ···················· | ···················· | ····················· | ···················· | ···················· | ···················· |
| E | 9616 hCoV-19/Wuhan/0125-A168/2020\|EPI_ISL_493159\|2020-0 | ···················· | ···················· | ····················· | ···················· | ···················· | ···················· |
| E | 9617 hCoV-19/Wuhan/0125-A170/2020\|EPI_ISL_493161\|2020-0 | ···················· | ···················· | ····················· | ···················· | ···················· | ···················· |
| E | 9618 hCoV-19/Wuhan/0125-A174/2020\|EPI_ISL_493163\|2020-0 | ···················· | ···················· | ····················· | ···················· | ···················· | ···················· |
| E | 9619 hCoV-19/Wuhan/0125-A182/2020\|EPI_ISL_493164\|2020-0 | ···················· | ···················· | ····················· | ···················· | ···················· | ···················· |
| E | 9620 hCoV-19/USA/CA-ALSR-1168/2020\|EPI_ISL_494410\|2020- | ···················· | ···················· | ····················· | ···················· | ···················· | ···················· |
| E | 9621 hCoV-19/USA/CA-ALSR-1169/2020\|EPI_ISL_494411\|2020- | ···················· | ···················· | ····················· | ···················· | ···················· | ···················· |
| E | 9622 hCoV-19/USA/CA-ALSR-1171/2020\|EPI_ISL_494412\|2020- | ···················· | ···················· | ····················· | ···················· | ···················· | ···················· |
| E | 9623 hCoV-19/USA/CA-ALSR-1172/2020\|EPI_ISL_494413\|2020- | ···················· | ···················· | ····················· | ···················· | ···················· | ···················· |
| E | 9624 hCoV-19/USA/CA-ALSR-1175/2020\|EPI_ISL_494414\|2020- | ···················· | ·A·················· | ····················· | ···················· | ···················· | ···················· |
| E | 9625 hCoV-19/USA/CA-ALSR-1177/2020\|EPI_ISL_494415\|2020- | ···················· | ·A·················· | ····················· | ···················· | ···················· | ···················· |
| E | 9626 hCoV-19/USA/CA-ALSR-1180/2020\|EPI_ISL_494416\|2020- | ···················· | ···················· | ····················· | ···················· | ···················· | ···················· |
| E | 9627 hCoV-19/USA/CA-ALSR-1181/2020\|EPI_ISL_494417\|2020- | ···················· | ···················· | ····················· | ···················· | ···················· | ···················· |
| E | 9628 hCoV-19/USA/CA-ALSR-1182/2020\|EPI_ISL_494418\|2020- | ···················· | ···················· | ····················· | ···················· | ···················· | ···················· |
| E | 9629 hCoV-19/USA/CA-ALSR-1183/2020\|EPI_ISL_494419\|2020- | ···················· | ···················· | ····················· | ···················· | ···················· | ···················· |
| E | 9630 hCoV-19/USA/CA-ALSR-1184/2020\|EPI_ISL_494420\|2020- | ···················· | ···················· | ····················· | ···················· | ···················· | ···················· |
| E | 9631 hCoV-19/USA/CA-ALSR-1185/2020\|EPI_ISL_494421\|2020- | ···················· | ···················· | ····················· | ···················· | ···················· | ···················· |
| E | 9632 hCoV-19/USA/CA-ALSR-1186/2020\|EPI_ISL_494422\|2020- | ···················· | ·A·················· | ····················· | ···················· | ···················· | ···················· |
| E | 9633 hCoV-19/USA/CA-ALSR-1187/2020\|EPI_ISL_494423\|2020- | ···················· | ·A·················· | ····················· | ···················· | ···················· | ···················· |
| E | 9634 hCoV-19/USA/CA-ALSR-1188/2020\|EPI_ISL_494424\|2020- | ···················· | ···················· | ····················· | ···················· | ···················· | ···················· |
| E | 9635 hCoV-19/USA/CA-ALSR-1189/2020\|EPI_ISL_494425\|2020- | ···················· | ·A·················· | ····················· | ···················· | ···················· | ···················· |
| E | 9636 hCoV-19/USA/CA-ALSR-1191/2020\|EPI_ISL_494427\|2020- | ···················· | ···················· | ····················· | ···················· | ···················· | ···················· |
| E | 9637 hCoV-19/USA/CA-ALSR-1192/2020\|EPI_ISL_494428\|2020- | ···················· | ·A·················· | ····················· | ···················· | ···················· | ···················· |
| E | 9638 hCoV-19/USA/CA-ALSR-1211/2020\|EPI_ISL_494441\|2020- | ···················· | ·A·················· | ····················· | ···················· | ···················· | ···················· |
| E | 9639 hCoV-19/USA/CA-ALSR-1214/2020\|EPI_ISL_494442\|2020- | ···················· | ·A·················· | ····················· | ···················· | ···················· | ···················· |
| E | 9640 hCoV-19/USA/CA-ALSR-1217/2020\|EPI_ISL_494443\|2020- | ···················· | ·A·················· | ····················· | ···················· | ···················· | ···················· |
| E | 9641 hCoV-19/USA/CA-ALSR-1219/2020\|EPI_ISL_494444\|2020- | ···················· | ·A·················· | ····················· | ···················· | ···················· | ···················· |
| E | 9642 hCoV-19/USA/CA-ALSR-1220/2020\|EPI_ISL_494445\|2020- | ···················· | ···················· | ····················· | ···················· | ···················· | ···················· |
| E | 9643 hCoV-19/USA/CA-ALSR-1222/2020\|EPI_ISL_494446\|2020- | ···················· | ·A·················· | ····················· | ···················· | ···················· | ···················· |
| E | 9644 hCoV-19/USA/CA-ALSR-1224/2020\|EPI_ISL_494448\|2020- | ···················· | ·A·················· | ····················· | ···················· | ···················· | ···················· |
| E | 9645 hCoV-19/USA/CA-ALSR-1225/2020\|EPI_ISL_494449\|2020- | ···················· | ·A·················· | ····················· | ···················· | ···················· | ···················· |
| E | 9646 hCoV-19/USA/CA-ALSR-1231/2020\|EPI_ISL_494452\|2020- | ···················· | ···················· | ····················· | ···················· | ···················· | ···················· |
| E | 9647 hCoV-19/USA/CA-ALSR-1239/2020\|EPI_ISL_494453\|2020- | ···················· | ·A·················· | ····················· | ···················· | ···················· | ···················· |
| E | 9648 hCoV-19/USA/CA-ALSR-1240/2020\|EPI_ISL_494454\|2020- | ···················· | ·A·················· | ····················· | ···················· | ···················· | ···················· |
| E | 9649 hCoV-19/USA/CA-ALSR-1242/2020\|EPI_ISL_494455\|2020- | ···················· | ·A·················· | ····················· | ···················· | ···················· | ···················· |
| E | 9650 hCoV-19/USA/CA-ALSR-1247/2020\|EPI_ISL_494456\|2020- | ···················· | ·A·················· | ····················· | ···················· | ···················· | ···················· |
| E | 9651 hCoV-19/USA/CA-ALSR-1248/2020\|EPI_ISL_494457\|2020- | ···················· | ·A·················· | ····················· | ···················· | ···················· | ···················· |
| E | 9652 hCoV-19/USA/CA-ALSR-1254/2020\|EPI_ISL_494458\|2020- | ···················· | ···················· | ····················· | ···················· | ···················· | ···················· |
| E | 9653 hCoV-19/USA/CA-ALSR-1255/2020\|EPI_ISL_494459\|2020- | ···················· | ·A·················· | ····················· | ···················· | ···················· | ···················· |
| E | 9654 hCoV-19/USA/CA-ALSR-1256/2020\|EPI_ISL_494460\|2020- | ···················· | ·A·················· | ····················· | ···················· | ···················· | ···················· |
| E | 9655 hCoV-19/USA/CA-ALSR-1260/2020\|EPI_ISL_494461\|2020- | ···················· | ·A·················· | ····················· | ···················· | ···················· | ···················· |
| E | 9656 hCoV-19/USA/CA-ALSR-1262/2020\|EPI_ISL_494463\|2020- | ···················· | ·A·················· | ····················· | ···················· | ···················· | ···················· |
| E | 9657 hCoV-19/USA/CA-ALSR-1263/2020\|EPI_ISL_494464\|2020- | ···················· | ·A·················· | ····················· | ···················· | ···················· | ···················· |
| E | 9658 hCoV-19/USA/CA-ALSR-1266/2020\|EPI_ISL_494466\|2020- | ···················· | ·A·················· | ····················· | ···················· | ···················· | ···················· |
| E | 9659 hCoV-19/USA/CA-ALSR-1267/2020\|EPI_ISL_494467\|2020- | ···················· | ·A·················· | ····················· | ···················· | ···················· | ···················· |
| E | 9660 hCoV-19/USA/CA-ALSR-1268/2020\|EPI_ISL_494468\|2020- | ···················· | ·A·················· | ····················· | ···················· | ···················· | ···················· |
| E | 9661 hCoV-19/USA/CA-ALSR-1272/2020\|EPI_ISL_494470\|2020- | ···················· | ·A·················· | ····················· | ···················· | ···················· | ···················· |
| E | 9662 hCoV-19/USA/CA-ALSR-1273/2020\|EPI_ISL_494471\|2020- | ···················· | ·A·················· | ····················· | ···················· | ···················· | ···················· |
| E | 9663 hCoV-19/USA/CA-ALSR-1274/2020\|EPI_ISL_494472\|2020- | ···················· | ···················· | ····················· | ···················· | ···················· | ···················· |
| E | 9664 hCoV-19/USA/CA-ALSR-1275/2020\|EPI_ISL_494473\|2020- | ···················· | ···················· | ····················· | ···················· | ···················· | ···················· |
| E | 9665 hCoV-19/USA/CA-ALSR-1276/2020\|EPI_ISL_494474\|2020- | ···················· | ·A·················· | ····················· | ···················· | ···················· | ···················· |
| E | 9666 hCoV-19/USA/CA-ALSR-1277/2020\|EPI_ISL_494475\|2020- | ···················· | ·A·················· | ····················· | ···················· | ···················· | ···················· |
| E | 9667 hCoV-19/USA/CA-ALSR-1279/2020\|EPI_ISL_494476\|2020- | ···················· | ·A·················· | ····················· | ···················· | ···················· | ···················· |
| E | 9668 hCoV-19/USA/CA-ALSR-1280/2020\|EPI_ISL_494477\|2020- | ···················· | ·A·················· | ····················· | ···················· | ···················· | ···················· |
| E | 9669 hCoV-19/USA/CA-ALSR-1282/2020\|EPI_ISL_494478\|2020- | ···················· | ·A·················· | ····················· | ···················· | ···················· | ···················· |
| E | 9671 hCoV-19/USA/CA-ALSR-1287/2020\|EPI_ISL_494480\|2020- | ···················· | ···················· | ····················· | ···················· | ···················· | ···················· |
| E | 9672 hCoV-19/USA/CA-ALSR-1288/2020\|EPI_ISL_494481\|2020- | ···················· | ·A·················· | ····················· | ···················· | ···················· | ···················· |
| E | 9673 hCoV-19/USA/CA-ALSR-1289/2020\|EPI_ISL_494482\|2020- | ···················· | ·A·················· | ····················· | ···················· | ···················· | ···················· |
| E | 9674 hCoV-19/USA/CA-ALSR-1290/2020\|EPI_ISL_494483\|2020- | ···················· | ·A·················· | ····················· | ···················· | ···················· | ···················· |
| E | 9675 hCoV-19/USA/CA-ALSR-1301/2020\|EPI_ISL_494485\|2020- | ···················· | ···················· | ····················· | ···················· | ···················· | ···················· |
| E | 9677 hCoV-19/USA/CA-ALSR-1303/2020\|EPI_ISL_494487\|2020- | ···················· | ···················· | ····················· | ···················· | ···················· | ···················· |
| E | 9678 hCoV-19/USA/CA-ALSR-1311/2020\|EPI_ISL_494489\|2020- | ···················· | ·A·················· | ····················· | ···················· | ···················· | ···················· |
| E | 9679 hCoV-19/USA/CA-ALSR-1314/2020\|EPI_ISL_494490\|2020- | ···················· | ·A·················· | ····················· | ···················· | ···················· | ···················· |
| E | 9680 hCoV-19/USA/CA-ALSR-1315/2020\|EPI_ISL_494491\|2020- | ···················· | ·A·················· | ····················· | ···················· | ···················· | ···················· |
| E | 9681 hCoV-19/USA/CA-ALSR-1320/2020\|EPI_ISL_494492\|2020- | ···················· | ···················· | ····················· | ···················· | ···················· | ···················· |
| E | 9682 hCoV-19/USA/CA-ALSR-1322/2020\|EPI_ISL_494493\|2020- | ···················· | ·A·················· | ····················· | ···················· | ···················· | ···················· |
| E | 9683 hCoV-19/USA/CA-ALSR-1324/2020\|EPI_ISL_494494\|2020- | ···················· | ·A·················· | ····················· | ···················· | ···················· | ···················· |
| E | 9684 hCoV-19/USA/CA-ALSR-1329/2020\|EPI_ISL_494495\|2020- | ···················· | ···················· | ····················· | ···················· | ···················· | ···················· |
| E | 9685 hCoV-19/USA/WA-S1290/2020\|EPI_ISL_495740\|2020-05-1 | ···················· | ···················· | ····················· | ···················· | ···················· | ···················· |
| E | 9686 hCoV-19/USA/WA-S1291/2020\|EPI_ISL_495741\|2020-05-1 | ···················· | ···················· | ····················· | ···················· | ···················· | ···················· |

| | | | | | | | |
|---|---|---|---|---|---|---|---|
| E | 9687 hCoV-19/USA/WA-S1292/2020\|EPI_ISL_495742\|2020-05-1 | ···················· | ···················· | ···················· | ···················· | ···················· | ···················· |
| E | 9688 hCoV-19/USA/WA-S1293/2020\|EPI_ISL_495743\|2020-05-1 | ···················· | ···················· | ···················· | ···················· | ···················· | ···················· |
| E | 9689 hCoV-19/USA/WA-S1294/2020\|EPI_ISL_495744\|2020-05-1 | ···················· | ···················· | ···················· | ···················· | ···················· | ···················· |
| E | 9690 hCoV-19/USA/WA-S1295/2020\|EPI_ISL_495745\|2020-05-1 | ···················· | ···················· | ···················· | ···················· | ···················· | ···················· |
| E | 9691 hCoV-19/USA/WA-S1296/2020\|EPI_ISL_495746\|2020-05-1 | ···················· | ···················· | ···················· | ···················· | ···················· | ···················· |
| E | 9692 hCoV-19/USA/WA-S1297/2020\|EPI_ISL_495747\|2020-05-1 | ···················· | ···················· | ···················· | ···················· | ···················· | ···················· |
| E | 9693 hCoV-19/USA/WA-S1298/2020\|EPI_ISL_495748\|2020-05-1 | ···················· | ···················· | ···················· | ···················· | ···················· | ···················· |
| E | 9694 hCoV-19/USA/WA-S1299/2020\|EPI_ISL_495749\|2020-05-1 | ···················· | ···················· | ···················· | ···················· | ···················· | ···················· |
| E | 9695 hCoV-19/USA/WA-S1300/2020\|EPI_ISL_495750\|2020-05-1 | ···················· | ···················· | ···················· | ···················· | ···················· | ···················· |
| E | 9696 hCoV-19/USA/WA-S1301/2020\|EPI_ISL_495751\|2020-05-1 | ···················· | ···················· | ···················· | ···················· | ···················· | ···················· |
| E | 9697 hCoV-19/USA/WA-S1302/2020\|EPI_ISL_495752\|2020-05-1 | ···················· | ···················· | ···················· | ···················· | ···················· | ···················· |
| E | 9698 hCoV-19/USA/WA-S1303/2020\|EPI_ISL_495753\|2020-05-1 | ···················· | ···················· | ···················· | ···················· | ···················· | ···················· |
| E | 9699 hCoV-19/USA/WA-S1304/2020\|EPI_ISL_495754\|2020-05-1 | ···················· | ···················· | ···················· | ···················· | ···················· | ···················· |
| E | 9700 hCoV-19/USA/WA-S1305/2020\|EPI_ISL_495755\|2020-05-1 | ···················· | ···················· | ···················· | ···················· | ···················· | ···················· |
| E | 9701 hCoV-19/USA/WA-S1306/2020\|EPI_ISL_495756\|2020-05-1 | ···················· | ···················· | ···················· | ···················· | ···················· | ···················· |
| E | 9702 hCoV-19/USA/WA-S1307/2020\|EPI_ISL_495757\|2020-05-1 | ···················· | ···················· | ···················· | ···················· | ···················· | ···················· |
| E | 9703 hCoV-19/USA/WA-S1308/2020\|EPI_ISL_495758\|2020-05-1 | ···················· | ···················· | ···················· | ···················· | ···················· | ···················· |
| E | 9704 hCoV-19/USA/WA-S1309/2020\|EPI_ISL_495759\|2020-05-1 | ···················· | ···················· | ···················· | ···················· | ···················· | ···················· |
| E | 9705 hCoV-19/USA/WA-S1310/2020\|EPI_ISL_495760\|2020-05-1 | ···················· | ···················· | ···················· | ···················· | ···················· | ···················· |
| E | 9706 hCoV-19/USA/WA-S1322/2020\|EPI_ISL_495772\|2020-05-1 | ···················· | ···················· | ···················· | ···················· | ···················· | ···················· |
| E | 9707 hCoV-19/USA/WA-S1323/2020\|EPI_ISL_495773\|2020-05-1 | ···················· | ···················· | ···················· | ···················· | ···················· | ···················· |
| E | 9708 hCoV-19/USA/WA-S1324/2020\|EPI_ISL_495774\|2020-05-1 | ···················· | ···················· | ···················· | ···················· | ···················· | ···················· |
| E | 9709 hCoV-19/USA/WA-S1325/2020\|EPI_ISL_495775\|2020-05-1 | ···················· | ···················· | ···················· | ···················· | ···················· | ···················· |
| E | 9710 hCoV-19/USA/WA-S1327/2020\|EPI_ISL_495777\|2020-05-1 | ···················· | ···················· | ···················· | ···················· | ···················· | ···················· |
| E | 9711 hCoV-19/USA/WA-S1328/2020\|EPI_ISL_495778\|2020-05-1 | ···················· | ···················· | ···················· | ···················· | ···················· | ···················· |
| E | 9712 hCoV-19/USA/WA-S1329/2020\|EPI_ISL_495779\|2020-05-1 | ···················· | ···················· | ···················· | ···················· | ···················· | ···················· |
| E | 9713 hCoV-19/USA/WA-S1334/2020\|EPI_ISL_495784\|2020-05-1 | ···················· | ···················· | ···················· | ···················· | ···················· | ···················· |
| E | 9714 hCoV-19/USA/WA-S1335/2020\|EPI_ISL_495785\|2020-05-1 | ···················· | ···················· | ···················· | ···················· | ···················· | ···················· |
| E | 9715 hCoV-19/USA/WA-S1336/2020\|EPI_ISL_495786\|2020-05-1 | ···················· | ···················· | ···················· | ···················· | ···················· | ···················· |
| E | 9716 hCoV-19/USA/WA-S1337/2020\|EPI_ISL_495787\|2020-05-0 | ···················· | ···················· | ···················· | ···················· | ···················· | ···················· |
| E | 9717 hCoV-19/USA/WA-S1338/2020\|EPI_ISL_495788\|2020-05-1 | ···················· | ···················· | ···················· | ···················· | ···················· | ···················· |
| E | 9718 hCoV-19/USA/WA-S1339/2020\|EPI_ISL_495789\|2020-05-1 | ···················· | ···················· | ···················· | ···················· | ···················· | ···················· |
| E | 9719 hCoV-19/USA/WA-S1340/2020\|EPI_ISL_495790\|2020-05-1 | ···················· | ···················· | ···················· | ···················· | ···················· | ···················· |
| E | 9720 hCoV-19/USA/WA-S1341/2020\|EPI_ISL_495791\|2020-05-1 | ···················· | ···················· | ···················· | ···················· | ···················· | ···················· |
| E | 9721 hCoV-19/USA/WA-S1342/2020\|EPI_ISL_495792\|2020-05-1 | ···················· | ···················· | ···················· | ···················· | ···················· | ···················· |
| E | 9722 hCoV-19/USA/WA-S1343/2020\|EPI_ISL_495793\|2020-05-1 | ···················· | ···················· | ···················· | ···················· | ···················· | ···················· |
| E | 9723 hCoV-19/USA/WA-S1344/2020\|EPI_ISL_495794\|2020-05-1 | ···················· | ···················· | ···················· | ···················· | ···················· | ···················· |
| E | 9724 hCoV-19/USA/WA-S1345/2020\|EPI_ISL_495795\|2020-05-1 | ···················· | ···················· | ···················· | ···················· | ···················· | ···················· |
| E | 9725 hCoV-19/USA/WA-S1346/2020\|EPI_ISL_495796\|2020-05-1 | ···················· | ···················· | ···················· | ···················· | ···················· | ···················· |
| E | 9726 hCoV-19/USA/WA-S1347/2020\|EPI_ISL_495797\|2020-05-1 | ···················· | ···················· | ···················· | ···················· | ···················· | ···················· |
| E | 9727 hCoV-19/USA/WA-S1348/2020\|EPI_ISL_495798\|2020-05-1 | ···················· | ···················· | ···················· | ···················· | ···················· | ···················· |
| E | 9728 hCoV-19/USA/WA-S1349/2020\|EPI_ISL_495799\|2020-05-1 | ···················· | ···················· | ···················· | ···················· | ···················· | ···················· |
| E | 9729 hCoV-19/India/OR-ILSCV27789/2020\|EPI_ISL_481124\|20 | ···················· | ···················· | ···················· | ···················· | ···················· | ···················· |
| E | 9730 hCoV-19/India/OR-ILSCV28062/2020\|EPI_ISL_481125\|20 | ···················· | ···················· | ···················· | ···················· | ···················· | ···················· |
| E | 9731 hCoV-19/India/OR-ILSCV28594/2020\|EPI_ISL_481126\|20 | ···················· | ···················· | ···················· | ···················· | ···················· | ···················· |
| E | 9732 hCoV-19/India/OR-ILSCV29181/2020\|EPI_ISL_481127\|20 | ···················· | ···················· | ···················· | ···················· | ···················· | ···················· |
| E | 9733 hCoV-19/India/OR-ILSCV29215/2020\|EPI_ISL_481128\|20 | ···················· | ···················· | ···················· | ···················· | ···················· | ···················· |
| E | 9734 hCoV-19/India/OR-ILSCV29457/2020\|EPI_ISL_481129\|20 | ···················· | ···················· | ···················· | ···················· | ···················· | ···················· |
| E | 9735 hCoV-19/India/OR-ILSCV29958/2020\|EPI_ISL_481131\|20 | ···················· | ···················· | ···················· | ···················· | ···················· | ···················· |
| E | 9736 hCoV-19/India/OR-ILSCV29959/2020\|EPI_ISL_481132\|20 | ···················· | ·A·················· | ···················· | ···················· | ···················· | ··A················· |
| E | 9737 hCoV-19/India/OR-ILSCV29960/2020\|EPI_ISL_481133\|20 | ···················· | ·A·················· | ···················· | ···················· | ···················· | ··A················· |
| E | 9738 hCoV-19/India/OR-ILSCV29961/2020\|EPI_ISL_481134\|20 | ···················· | ·A·················· | ···················· | ···················· | ···················· | ··A················· |
| E | 9739 hCoV-19/India/OR-ILSCV29962/2020\|EPI_ISL_481135\|20 | ···················· | ·A·················· | ···················· | ···················· | ···················· | ··A················· |
| E | 9740 hCoV-19/India/OR-ILSCV29971/2020\|EPI_ISL_481136\|20 | ···················· | ···················· | ···················· | ···················· | ···················· | ···················· |
| E | 9741 hCoV-19/India/OR-ILSCV29997/2020\|EPI_ISL_481137\|20 | ···················· | ···················· | ···················· | ···················· | ···················· | ···················· |
| E | 9742 hCoV-19/India/OR-ILSCV30115/2020\|EPI_ISL_481138\|20 | ···················· | ···················· | ···················· | ···················· | ···················· | ···················· |
| E | 9743 hCoV-19/India/OR-ILSCV30140/2020\|EPI_ISL_481139\|20 | ···················· | ···················· | ···················· | ···················· | ···················· | ···················· |
| E | 9744 hCoV-19/India/OR-ILSCV30141/2020\|EPI_ISL_481140\|20 | ···················· | ···················· | ···················· | ···················· | ···················· | ···················· |
| E | 9745 hCoV-19/India/OR-ILSCV30163/2020\|EPI_ISL_481141\|20 | ···················· | ···················· | ···················· | ···················· | ···················· | ···················· |
| E | 9746 hCoV-19/USA/OR-PROV-139/2020\|EPI_ISL_482400\|2020-0 | ···················· | ···················· | ···················· | ···················· | ···················· | ···················· |
| E | 9747 hCoV-19/USA/OR-PROV-143/2020\|EPI_ISL_482401\|2020-0 | ···················· | ···················· | ···················· | ···················· | ···················· | ···················· |
| E | 9748 hCoV-19/USA/OR-PROV-144/2020\|EPI_ISL_482402\|2020-0 | ···················· | ···················· | ···················· | ···················· | ···················· | ···················· |
| E | 9749 hCoV-19/USA/OR-PROV-145/2020\|EPI_ISL_482403\|2020-0 | ···················· | ···················· | ···················· | ···················· | ···················· | ···················· |
| E | 9750 hCoV-19/USA/OR-PROV-146/2020\|EPI_ISL_482404\|2020-0 | ···················· | ···················· | ···················· | ···················· | ···················· | ···················· |
| E | 9751 hCoV-19/USA/OR-PROV-147/2020\|EPI_ISL_482405\|2020-0 | ···················· | ···················· | ···················· | ···················· | ···················· | ···················· |
| E | 9752 hCoV-19/USA/OR-PROV-148/2020\|EPI_ISL_482406\|2020-0 | ···················· | ···················· | ···················· | ···················· | ···················· | ···················· |
| E | 9753 hCoV-19/USA/OR-PROV-151/2020\|EPI_ISL_482407\|2020-0 | ···················· | ···················· | ···················· | ···················· | ···················· | ···················· |
| E | 9754 hCoV-19/USA/OR-PROV-153/2020\|EPI_ISL_482408\|2020-0 | ···················· | ···················· | ···················· | ···················· | ···················· | ···················· |
| E | 9755 hCoV-19/USA/OR-PROV-154/2020\|EPI_ISL_482409\|2020-0 | ···················· | ···················· | ···················· | ···················· | ···················· | ···················· |
| E | 9756 hCoV-19/USA/OR-PROV-155/2020\|EPI_ISL_482410\|2020-0 | ···················· | ···················· | ···················· | ···················· | ···················· | ···················· |
| E | 9757 hCoV-19/USA/OR-PROV-156/2020\|EPI_ISL_482411\|2020-0 | ···················· | ···················· | ···················· | ···················· | ···················· | ···················· |
| E | 9758 hCoV-19/USA/OR-PROV-157/2020\|EPI_ISL_482412\|2020-0 | ···················· | ···················· | ···················· | ···················· | ···················· | ···················· |
| E | 9759 hCoV-19/USA/OR-PROV-158/2020\|EPI_ISL_482413\|2020-0 | ···················· | ···················· | ···················· | ···················· | ···················· | ···················· |
| E | 9760 hCoV-19/USA/OR-PROV-159/2020\|EPI_ISL_482414\|2020-0 | ···················· | ···················· | ···················· | ···················· | ···················· | ···················· |
| E | 9761 hCoV-19/USA/OR-PROV-161/2020\|EPI_ISL_482415\|2020-0 | ···················· | ···················· | ···················· | ···················· | ···················· | ···················· |
| E | 9762 hCoV-19/USA/OR-PROV-163/2020\|EPI_ISL_482416\|2020-0 | ···················· | ···················· | ···················· | ···················· | ···················· | ···················· |
| E | 9763 hCoV-19/USA/OR-PROV-165/2020\|EPI_ISL_482417\|2020-0 | ···················· | ···················· | ···················· | ···················· | ···················· | ···················· |
| E | 9764 hCoV-19/USA/OR-PROV-167/2020\|EPI_ISL_482418\|2020-0 | ···················· | ···················· | ···················· | ···················· | ···················· | ···················· |
| E | 9765 hCoV-19/USA/OR-PROV-168/2020\|EPI_ISL_482419\|2020-0 | ···················· | ···················· | ···················· | ···················· | ···················· | ···················· |
| E | 9766 hCoV-19/USA/OR-PROV-169/2020\|EPI_ISL_482420\|2020-0 | ···················· | ···················· | ···················· | ···················· | ···················· | ···················· |

| | | | | | | | | |
|---|---|---|---|---|---|---|---|---|
| E | 9767 | hCoV-19/USA/OR-PROV-170/2020\|EPI_ISL_482421\|2020-0 | ···················· | ···················· | ···················· | ···················· | ···················· | ···················· |
| E | 9768 | hCoV-19/USA/OR-PROV-171/2020\|EPI_ISL_482422\|2020-0 | ···················· | ···················· | ···················· | ···················· | ···················· | ···················· |
| E | 9769 | hCoV-19/USA/OR-PROV-173/2020\|EPI_ISL_482423\|2020-0 | ···················· | ···················· | ···················· | ···················· | ···················· | ···················· |
| E | 9770 | hCoV-19/USA/OR-PROV-174/2020\|EPI_ISL_482424\|2020-0 | ···················· | ···················· | ···················· | ···················· | ···················· | ···················· |
| E | 9771 | hCoV-19/USA/OR-PROV-175/2020\|EPI_ISL_482425\|2020-0 | ···················· | ···················· | ···················· | ···················· | ···················· | ···················· |
| E | 9772 | hCoV-19/USA/OR-PROV-176/2020\|EPI_ISL_482426\|2020-0 | ···················· | ···················· | ···················· | ···················· | ···················· | ···················· |
| E | 9773 | hCoV-19/USA/OR-PROV-178/2020\|EPI_ISL_482427\|2020-0 | ···················· | ···················· | ···················· | ···················· | ···················· | ···················· |
| E | 9774 | hCoV-19/USA/OR-PROV-179/2020\|EPI_ISL_482428\|2020-0 | ···················· | ···················· | ···················· | ···················· | ···················· | ···················· |
| E | 9775 | hCoV-19/USA/OR-PROV-180/2020\|EPI_ISL_482429\|2020-0 | ···················· | ···················· | ···················· | ···················· | ···················· | ···················· |
| E | 9776 | hCoV-19/USA/OR-PROV-220/2020\|EPI_ISL_482455\|2020-0 | ···················· | ···················· | ···················· | ···················· | ···················· | ···················· |
| E | 9777 | hCoV-19/USA/OR-PROV-221/2020\|EPI_ISL_482456\|2020-0 | ···················· | ···················· | ···················· | ···················· | ···················· | ···················· |
| E | 9778 | hCoV-19/USA/OR-PROV-222/2020\|EPI_ISL_482457\|2020-0 | ···················· | ···················· | ···················· | ···················· | ···················· | ···················· |
| E | 9779 | hCoV-19/USA/OR-PROV-223/2020\|EPI_ISL_482458\|2020-0 | ···················· | ···················· | ···················· | ···················· | ···················· | ···················· |
| E | 9780 | hCoV-19/USA/OR-PROV-224/2020\|EPI_ISL_482459\|2020-0 | ···················· | ···················· | ···················· | ···················· | ···················· | ···················· |
| E | 9781 | hCoV-19/USA/OR-PROV-227/2020\|EPI_ISL_482462\|2020-0 | ···················· | ···················· | ···················· | ···················· | ···················· | ···················· |
| E | 9782 | hCoV-19/USA/OR-PROV-228/2020\|EPI_ISL_482463\|2020-0 | ···················· | ···················· | ···················· | ···················· | ···················· | ···················· |
| E | 9783 | hCoV-19/USA/OR-PROV-229/2020\|EPI_ISL_482464\|2020-0 | ···················· | ···················· | ···················· | ···················· | ···················· | ···················· |
| E | 9784 | hCoV-19/USA/OR-PROV-230/2020\|EPI_ISL_482465\|2020-0 | ···················· | ···················· | ···················· | ···················· | ···················· | ···················· |
| E | 9785 | hCoV-19/USA/OR-PROV-231/2020\|EPI_ISL_482466\|2020-0 | ···················· | ···················· | ···················· | ···················· | ···················· | ···················· |
| E | 9786 | hCoV-19/USA/OR-PROV-232/2020\|EPI_ISL_482467\|2020-0 | ···················· | ···················· | ···················· | ···················· | ···················· | ···················· |
| E | 9787 | hCoV-19/Peru/LIM-01-2/2020\|EPI_ISL_482468\|2020-03- | ···················· | ···················· | ···················· | ···················· | ···················· | ···················· |
| E | 9788 | hCoV-19/Canada/NS_40/2020\|EPI_ISL_482469\|2020-03-0 | ···················· | ···················· | ···················· | ···················· | ···················· | ···················· |
| E | 9789 | hCoV-19/Canada/NS_42/2020\|EPI_ISL_482470\|2020-03-1 | ···················· | ···················· | ···················· | ···················· | ···················· | ···················· |
| E | 9790 | hCoV-19/Canada/NB_54/2020\|EPI_ISL_482471\|2020-03-1 | ···················· | ···················· | ···················· | ···················· | ···················· | ···················· |
| E | 9791 | hCoV-19/Canada/NB_55/2020\|EPI_ISL_482472\|2020-03-1 | ···················· | ···················· | ···················· | ···················· | ···················· | ···················· |
| E | 9792 | hCoV-19/Bangladesh/BCSIR-NILMRC-203/2020\|EPI_ISL_4 | ···················· | ···················· | ···················· | ···················· | ···················· | ···················· |
| E | 9793 | hCoV-19/Bangladesh/BCSIR-NILMRC-216/2020\|EPI_ISL_4 | ···················· | ·A·················· | ···················· | ···················· | ···················· | ··A················· |
| E | 9794 | hCoV-19/Israel/n14269/2020\|EPI_ISL_483704\|2020-03- | ···················· | ···················· | ···················· | ···················· | ···················· | ···················· |
| E | 9795 | hCoV-19/Bangladesh/BCSIR-NILMRC-218/2020\|EPI_ISL_4 | ···················· | ···················· | ···················· | ···················· | ···················· | ···················· |
| E | 9796 | hCoV-19/Bangladesh/BCSIR-NILMRC-219/2020\|EPI_ISL_4 | ···················· | ···················· | ···················· | ···················· | ···················· | ···················· |
| E | 9797 | hCoV-19/Israel/n14268/2020\|EPI_ISL_483708\|2020-03- | ···················· | ···················· | ···················· | ···················· | ···················· | ···················· |
| E | 9798 | hCoV-19/Israel/n14279/2020\|EPI_ISL_483709\|2020-03- | ···················· | ···················· | ···················· | ···················· | ···················· | ···················· |
| E | 9799 | hCoV-19/Bangladesh/BCSIR-NILMRC-220/2020\|EPI_ISL_4 | ···················· | ·A·················· | ···················· | ···················· | ···················· | ··A················· |
| E | 9800 | hCoV-19/Israel/n14285/2020\|EPI_ISL_483711\|2020-03- | ···················· | ···················· | ···················· | ···················· | ···················· | ···················· |
| E | 9801 | hCoV-19/Israel/n14292/2020\|EPI_ISL_483712\|2020-04- | ···················· | ···················· | ···················· | ···················· | ···················· | ···················· |
| E | 9802 | hCoV-19/Israel/n14293/2020\|EPI_ISL_483713\|2020-04- | ···················· | ···················· | ···················· | ···················· | ···················· | ···················· |
| E | 9803 | hCoV-19/Israel/n14270/2020\|EPI_ISL_483715\|2020-03- | ···················· | ···················· | ···················· | ···················· | ···················· | ···················· |
| E | 9804 | hCoV-19/Israel/n14271/2020\|EPI_ISL_483717\|2020-03- | ···················· | ···················· | ···················· | ···················· | ···················· | ···················· |
| E | 9805 | hCoV-19/Israel/n14289/2020\|EPI_ISL_483725\|2020-04- | ···················· | ···················· | ···················· | ···················· | ···················· | ···················· |
| E | 9807 | hCoV-19/South Africa/KRISP-0318/2020\|EPI_ISL_48729 | ···················· | ···················· | ···················· | ···················· | ···················· | ···················· |
| E | 9808 | hCoV-19/USA/UT-02512/2020\|EPI_ISL_493100\|2020-04-2 | ···················· | ···················· | ···················· | ···················· | ···················· | ···················· |
| E | 9809 | hCoV-19/USA/UT-02514/2020\|EPI_ISL_493102\|2020-04-2 | ···················· | ···················· | ···················· | ···················· | ···················· | ···················· |
| E | 9810 | hCoV-19/USA/UT-02516/2020\|EPI_ISL_493104\|2020-04-2 | ···················· | ···················· | ···················· | ···················· | ···················· | ···················· |
| E | 9811 | hCoV-19/USA/UT-02519/2020\|EPI_ISL_493107\|2020-04-2 | ···················· | ···················· | ···················· | ···················· | ···················· | ···················· |
| E | 9812 | hCoV-19/USA/CA-ALSR-1157/2020\|EPI_ISL_494400\|2020- | ···················· | ···················· | ···················· | ···················· | ···················· | ···················· |
| E | 9813 | hCoV-19/USA/CA-ALSR-1158/2020\|EPI_ISL_494401\|2020- | ···················· | ·A·················· | ···················· | ···················· | ···················· | ···················· |
| E | 9814 | hCoV-19/USA/CA-ALSR-1159/2020\|EPI_ISL_494402\|2020- | ···················· | ···················· | ···················· | ···················· | ···················· | ···················· |
| E | 9815 | hCoV-19/USA/CA-ALSR-1160/2020\|EPI_ISL_494403\|2020- | ···················· | ·A·················· | ···················· | ···················· | ···················· | ···················· |
| E | 9816 | hCoV-19/USA/CA-ALSR-1161/2020\|EPI_ISL_494404\|2020- | ···················· | ···················· | ···················· | ···················· | ···················· | ···················· |
| E | 9817 | hCoV-19/USA/CA-ALSR-1162/2020\|EPI_ISL_494405\|2020- | ···················· | ···················· | ···················· | ···················· | ···················· | ···················· |
| E | 9818 | hCoV-19/USA/CA-ALSR-1164/2020\|EPI_ISL_494406\|2020- | ···················· | ···················· | ···················· | ···················· | ···················· | ···················· |
| E | 9819 | hCoV-19/USA/CA-ALSR-1165/2020\|EPI_ISL_494407\|2020- | ···················· | ···················· | ···················· | ···················· | ···················· | ···················· |
| E | 9820 | hCoV-19/USA/CA-ALSR-1166/2020\|EPI_ISL_494408\|2020- | ···················· | ···················· | ···················· | ···················· | ···················· | ···················· |
| E | 9821 | hCoV-19/USA/CA-ALSR-1167/2020\|EPI_ISL_494409\|2020- | ···················· | ···················· | ···················· | ···················· | ···················· | ···················· |
| E | 9822 | hCoV-19/USA/CA-ALSR-1196/2020\|EPI_ISL_494430\|2020- | ···················· | ·A·················· | ···················· | ···················· | ···················· | ···················· |
| E | 9823 | hCoV-19/USA/CA-ALSR-1197/2020\|EPI_ISL_494431\|2020- | ···················· | ·A·················· | ···················· | ···················· | ···················· | ···················· |
| E | 9824 | hCoV-19/USA/CA-ALSR-1200/2020\|EPI_ISL_494432\|2020- | ···················· | ·A·················· | ···················· | ···················· | ···················· | ···················· |
| E | 9825 | hCoV-19/USA/CA-ALSR-1201/2020\|EPI_ISL_494433\|2020- | ···················· | ·A·················· | ···················· | ···················· | ···················· | ···················· |
| E | 9826 | hCoV-19/USA/CA-ALSR-1202/2020\|EPI_ISL_494434\|2020- | ···················· | ···················· | ···················· | ···················· | ···················· | ···················· |
| E | 9827 | hCoV-19/USA/CA-ALSR-1203/2020\|EPI_ISL_494435\|2020- | ···················· | ·A·················· | ···················· | ···················· | ···················· | ···················· |
| E | 9828 | hCoV-19/USA/CA-ALSR-1204/2020\|EPI_ISL_494436\|2020- | ···················· | ·A·················· | ···················· | ···················· | ···················· | ···················· |
| E | 9829 | hCoV-19/USA/CA-ALSR-1207/2020\|EPI_ISL_494437\|2020- | ···················· | ·A·················· | ···················· | ···················· | ···················· | ···················· |
| E | 9830 | hCoV-19/USA/CA-ALSR-1208/2020\|EPI_ISL_494438\|2020- | ···················· | ·A·················· | ···················· | ···················· | ···················· | ···················· |
| E | 9831 | hCoV-19/USA/CA-ALSR-1209/2020\|EPI_ISL_494439\|2020- | ···················· | ·A·················· | ···················· | ···················· | ···················· | ···················· |
| E | 9832 | hCoV-19/USA/CA-ALSR-1210/2020\|EPI_ISL_494440\|2020- | ···················· | ·A·················· | ···················· | ···················· | ···················· | ···················· |
| E | 9833 | hCoV-19/USA/CA-ALSR-1228/2020\|EPI_ISL_494450\|2020- | ···················· | ·A·················· | ···················· | ···················· | ···················· | ···················· |
| E | 9834 | hCoV-19/USA/CA-ALSR-1229/2020\|EPI_ISL_494451\|2020- | ···················· | ·A·················· | ···················· | ···················· | ···················· | ···················· |
| E | 9835 | hCoV-19/USA/WA-S1250/2020\|EPI_ISL_495700\|2020-05-1 | ···················· | ···················· | ···················· | ···················· | ···················· | ···················· |
| E | 9836 | hCoV-19/USA/WA-S1251/2020\|EPI_ISL_495701\|2020-05-1 | ···················· | ···················· | ···················· | ···················· | ···················· | ···················· |
| E | 9837 | hCoV-19/USA/WA-S1253/2020\|EPI_ISL_495703\|2020-05-1 | ···················· | ···················· | ···················· | ···················· | ···················· | ···················· |
| E | 9838 | hCoV-19/USA/WA-S1254/2020\|EPI_ISL_495704\|2020-05-1 | ···················· | ···················· | ···················· | ···················· | ···················· | ···················· |
| E | 9839 | hCoV-19/USA/WA-S1255/2020\|EPI_ISL_495705\|2020-05-1 | ···················· | ···················· | ···················· | ···················· | ···················· | ···················· |
| E | 9840 | hCoV-19/USA/WA-S1256/2020\|EPI_ISL_495706\|2020-05-1 | ···················· | ···················· | ···················· | ···················· | ···················· | ···················· |
| E | 9841 | hCoV-19/USA/WA-S1257/2020\|EPI_ISL_495707\|2020-05-1 | ···················· | ···················· | ···················· | ···················· | ···················· | ···················· |
| E | 9842 | hCoV-19/USA/WA-S1258/2020\|EPI_ISL_495708\|2020-05-1 | ···················· | ···················· | ···················· | ···················· | ···················· | ···················· |
| E | 9843 | hCoV-19/USA/WA-S1259/2020\|EPI_ISL_495709\|2020-05-1 | ···················· | ···················· | ···················· | ···················· | ···················· | ···················· |
| E | 9844 | hCoV-19/USA/WA-S1260/2020\|EPI_ISL_495710\|2020-05-1 | ···················· | ···················· | ···················· | ···················· | ···················· | ···················· |
| E | 9845 | hCoV-19/USA/WA-S1261/2020\|EPI_ISL_495711\|2020-05-1 | ···················· | ···················· | ···················· | ···················· | ···················· | ···················· |
| E | 9846 | hCoV-19/USA/WA-S1262/2020\|EPI_ISL_495712\|2020-05-1 | ···················· | ···················· | ···················· | ···················· | ···················· | ···················· |
| E | 9847 | hCoV-19/USA/WA-S1263/2020\|EPI_ISL_495713\|2020-05-1 | ···················· | ···················· | ···················· | ···················· | ···················· | ···················· |

| | | | | | | | | |
|---|---|---|---|---|---|---|---|---|
| E | 9848 | hCoV-19/USA/WA-S1264/2020\|EPI_ISL_495714\|2020-05-1 | .................... | .................... | ..................... | .................... | ............T....... | .................... |
| E | 9849 | hCoV-19/USA/WA-S1265/2020\|EPI_ISL_495715\|2020-05-1 | .................... | .................... | ..................... | .................... | ............T....... | .................... |
| E | 9850 | hCoV-19/USA/WA-S1266/2020\|EPI_ISL_495716\|2020-05-1 | .................... | .................... | ..................... | .................... | .................... | .................... |
| E | 9851 | hCoV-19/USA/WA-S1267/2020\|EPI_ISL_495717\|2020-05-0 | .................... | .................... | ..................... | .................... | .................... | .................... |
| E | 9852 | hCoV-19/USA/WA-S1268/2020\|EPI_ISL_495718\|2020-05-1 | .................... | .................... | ..................... | .................... | .................... | .................... |
| E | 9853 | hCoV-19/USA/WA-S1269/2020\|EPI_ISL_495719\|2020-05-1 | .................... | .................... | ..................... | .................... | .................... | .................... |
| E | 9854 | hCoV-19/USA/WA-S1270/2020\|EPI_ISL_495720\|2020-05-1 | .................... | .................... | ..................... | .................... | .................... | .................... |
| E | 9855 | hCoV-19/USA/WA-S1271/2020\|EPI_ISL_495721\|2020-05-1 | .................... | .................... | ..................... | .................... | .................... | .................... |
| E | 9856 | hCoV-19/USA/WA-S1272/2020\|EPI_ISL_495722\|2020-05-1 | .................... | .................... | ..................... | .................... | .................... | .................... |
| E | 9857 | hCoV-19/USA/WA-S1273/2020\|EPI_ISL_495723\|2020-05-1 | .................... | .................... | ..................... | .................... | .................... | .................... |
| E | 9858 | hCoV-19/USA/WA-S1274/2020\|EPI_ISL_495724\|2020-05-1 | .................... | .................... | ..................... | .................... | .................... | .................... |
| E | 9859 | hCoV-19/USA/WA-S1275/2020\|EPI_ISL_495725\|2020-05-1 | .................... | .................... | ..................... | .................... | .................... | .................... |
| E | 9860 | hCoV-19/USA/WA-S1276/2020\|EPI_ISL_495726\|2020-05-1 | .................... | .................... | ..................... | .................... | .................... | .................... |
| E | 9861 | hCoV-19/USA/WA-S1277/2020\|EPI_ISL_495727\|2020-05-1 | .................... | .................... | ..................... | .................... | .................... | .................... |
| E | 9862 | hCoV-19/USA/WA-S1278/2020\|EPI_ISL_495728\|2020-05-1 | .................... | .................... | ..................... | .................... | .................... | .................... |
| E | 9863 | hCoV-19/USA/WA-S1279/2020\|EPI_ISL_495729\|2020-05-1 | .................... | .A.................. | ..................... | .................... | .................... | .................... |
| E | 9864 | hCoV-19/USA/WA-S1280/2020\|EPI_ISL_495730\|2020-05-1 | .................... | .................... | ..................... | .................... | .................... | .................... |
| E | 9865 | hCoV-19/USA/WA-S1281/2020\|EPI_ISL_495731\|2020-05-1 | .................... | .................... | ..................... | .................... | .................... | .................... |
| E | 9866 | hCoV-19/USA/WA-S1282/2020\|EPI_ISL_495732\|2020-05-1 | .................... | .................... | ..................... | .................... | .................... | .................... |
| E | 9867 | hCoV-19/USA/WA-S1283/2020\|EPI_ISL_495733\|2020-05-1 | .................... | .................... | ..................... | .................... | .................... | .................... |
| E | 9868 | hCoV-19/USA/WA-S1284/2020\|EPI_ISL_495734\|2020-05-1 | .................... | .................... | ..................... | .................... | .................... | .................... |
| E | 9869 | hCoV-19/USA/WA-S1285/2020\|EPI_ISL_495735\|2020-05-1 | .................... | .................... | ..................... | .................... | .................... | .................... |
| E | 9870 | hCoV-19/USA/WA-S1286/2020\|EPI_ISL_495736\|2020-05-1 | .................... | .................... | ..................... | .................... | .................... | .................... |
| E | 9871 | hCoV-19/USA/WA-S1287/2020\|EPI_ISL_495737\|2020-05-1 | .................... | .................... | ..................... | .................... | .................... | .................... |
| E | 9872 | hCoV-19/USA/WA-S1288/2020\|EPI_ISL_495738\|2020-05-1 | .................... | .................... | ..................... | .................... | .................... | .................... |
| E | 9873 | hCoV-19/USA/WA-S1289/2020\|EPI_ISL_495739\|2020-05-1 | .................... | .................... | ..................... | .................... | .................... | .................... |
| E | 9874 | hCoV-19/USA/WA-S1311/2020\|EPI_ISL_495761\|2020-05-1 | .................... | .................... | ..................... | .................... | .................... | .................... |
| E | 9875 | hCoV-19/USA/WA-S1312/2020\|EPI_ISL_495762\|2020-05-1 | .................... | .................... | ..................... | .................... | .................... | .................... |
| E | 9876 | hCoV-19/USA/WA-S1313/2020\|EPI_ISL_495763\|2020-05-1 | .................... | .................... | ..................... | .................... | .................... | .................... |
| E | 9877 | hCoV-19/USA/WA-S1314/2020\|EPI_ISL_495764\|2020-05-1 | .................... | .................... | ..................... | .................... | .................... | .................... |
| E | 9878 | hCoV-19/USA/WA-S1315/2020\|EPI_ISL_495765\|2020-05-1 | .................... | .................... | ..................... | .................... | .................... | .................... |
| E | 9879 | hCoV-19/USA/WA-S1316/2020\|EPI_ISL_495766\|2020-05-1 | .................... | .................... | ..................... | .................... | .................... | .................... |
| E | 9880 | hCoV-19/USA/WA-S1318/2020\|EPI_ISL_495768\|2020-05-1 | .................... | .................... | ..................... | .................... | .................... | .................... |
| E | 9881 | hCoV-19/USA/WA-S1319/2020\|EPI_ISL_495769\|2020-05-1 | .................... | .................... | ..................... | .................... | .................... | .................... |
| E | 9882 | hCoV-19/USA/WA-S1321/2020\|EPI_ISL_495771\|2020-05-1 | .................... | .................... | ..................... | .................... | .................... | .................... |
| E | 9883 | hCoV-19/USA/WA-S1330/2020\|EPI_ISL_495780\|2020-05-1 | .................... | .................... | ..................... | .................... | .................... | .................... |
| E | 9884 | hCoV-19/USA/WA-S1331/2020\|EPI_ISL_495781\|2020-05-1 | .................... | .................... | ..................... | .................... | .................... | .................... |
| E | 9885 | hCoV-19/USA/WA-S1332/2020\|EPI_ISL_495782\|2020-05-1 | .................... | .................... | ..................... | .................... | .................... | .................... |
| F | 1 | hCoV-19/USA/WA-S2570/2020\|EPI_ISL_497202\|2020-06-1 | .................... | .................... | ..................... | .................... | .................... | .................... |
| F | 2 | hCoV-19/USA/WA-S2571/2020\|EPI_ISL_497203\|2020-06-1 | .................... | .................... | ..................... | .................... | .................... | .................... |
| F | 3 | hCoV-19/USA/WA-S2573/2020\|EPI_ISL_497205\|2020-06-1 | .................... | .................... | ..................... | .................... | .................... | .................... |
| F | 4 | hCoV-19/USA/WA-S2574/2020\|EPI_ISL_497206\|2020-06-1 | .................... | .................... | ..................... | .................... | .................... | .................... |
| F | 5 | hCoV-19/USA/WA-S2575/2020\|EPI_ISL_497207\|2020-06-1 | .................... | .................... | ..................... | .................... | .................... | .................... |
| F | 6 | hCoV-19/USA/WA-S2576/2020\|EPI_ISL_497208\|2020-06-1 | .................... | .................... | ..................... | .................... | .................... | .................... |
| F | 7 | hCoV-19/USA/WA-S2577/2020\|EPI_ISL_497209\|2020-06-1 | .................... | .................... | ..................... | .................... | .................... | .................... |
| F | 8 | hCoV-19/USA/WA-S2578/2020\|EPI_ISL_497210\|2020-06-1 | .................... | .................... | ..................... | .................... | .................... | .................... |
| F | 9 | hCoV-19/USA/WA-S2579/2020\|EPI_ISL_497211\|2020-06-1 | .................... | .................... | ..................... | .................... | .................... | .................... |
| F | 10 | hCoV-19/USA/WA-S2580/2020\|EPI_ISL_497212\|2020-06-1 | .................... | .................... | ..................... | .................... | .................... | .................... |
| F | 11 | hCoV-19/USA/WA-S2581/2020\|EPI_ISL_497213\|2020-06-1 | .................... | .................... | ..................... | .................... | .................... | .................... |
| F | 12 | hCoV-19/USA/WA-S2582/2020\|EPI_ISL_497214\|2020-06-1 | .................... | .................... | ..................... | .................... | .................... | .................... |
| F | 13 | hCoV-19/USA/WA-S2583/2020\|EPI_ISL_497215\|2020-06-1 | .................... | .................... | ..................... | .................... | .................... | .................... |
| F | 14 | hCoV-19/USA/WA-S2584/2020\|EPI_ISL_497216\|2020-06-1 | .................... | .A.................. | ..................... | .................... | .................... | .................... |
| F | 15 | hCoV-19/USA/WA-S2585/2020\|EPI_ISL_497217\|2020-06-1 | .................... | .................... | ..................... | .................... | .................... | .................... |
| F | 16 | hCoV-19/USA/WA-S2586/2020\|EPI_ISL_497218\|2020-06-1 | .................... | .................... | ..................... | .................... | .................... | .................... |
| F | 17 | hCoV-19/USA/WA-S2587/2020\|EPI_ISL_497219\|2020-06-1 | .................... | .................... | ..................... | .................... | .................... | .................... |
| F | 18 | hCoV-19/USA/WA-S2588/2020\|EPI_ISL_497220\|2020-06-1 | .................... | .................... | ..................... | .................... | .................... | .................... |
| F | 19 | hCoV-19/USA/WA-S2589/2020\|EPI_ISL_497221\|2020-06-1 | .................... | .................... | ..................... | .................... | .................... | .................... |
| F | 20 | hCoV-19/USA/WA-S2590/2020\|EPI_ISL_497222\|2020-06-1 | .................... | .................... | ..................... | .................... | .................... | .................... |
| F | 21 | hCoV-19/USA/WA-S2591/2020\|EPI_ISL_497223\|2020-06-1 | .................... | .................... | ..................... | .................... | .................... | .................... |
| F | 22 | hCoV-19/USA/WA-S2614/2020\|EPI_ISL_497246\|2020-06-1 | .................... | .................... | ..................... | .................... | .................... | .................... |
| F | 23 | hCoV-19/USA/WA-S2615/2020\|EPI_ISL_497247\|2020-06-1 | .................... | .................... | ..................... | .................... | .................... | .................... |
| F | 24 | hCoV-19/USA/WA-S2616/2020\|EPI_ISL_497248\|2020-06-1 | .................... | .................... | ..................... | .................... | .................... | .................... |
| F | 25 | hCoV-19/USA/WA-S2617/2020\|EPI_ISL_497249\|2020-06-2 | .................... | .................... | ..................... | .................... | .................... | .................... |
| F | 26 | hCoV-19/USA/WA-S2618/2020\|EPI_ISL_497250\|2020-06-1 | .................... | .................... | ..................... | .................... | .................... | .................... |
| F | 27 | hCoV-19/USA/WA-S2619/2020\|EPI_ISL_497251\|2020-06-1 | .................... | .................... | ..................... | .................... | .................... | .................... |
| F | 28 | hCoV-19/USA/WA-S2620/2020\|EPI_ISL_497252\|2020-06-1 | .................... | .................... | ..................... | .................... | .................... | .................... |
| F | 29 | hCoV-19/USA/WA-S2621/2020\|EPI_ISL_497253\|2020-06-1 | .................... | .................... | ..................... | .................... | .................... | .................... |
| F | 30 | hCoV-19/USA/WA-S2622/2020\|EPI_ISL_497254\|2020-06-1 | .................... | .................... | ..................... | .................... | .................... | .................... |
| F | 31 | hCoV-19/USA/WA-S2623/2020\|EPI_ISL_497255\|2020-06-1 | .................... | .................... | ..................... | .................... | .................... | .................... |
| F | 32 | hCoV-19/USA/WA-S2624/2020\|EPI_ISL_497256\|2020-06-1 | .................... | .................... | ..................... | .................... | .................... | .................... |
| F | 33 | hCoV-19/USA/WA-S2625/2020\|EPI_ISL_497257\|2020-06-1 | .................... | .................... | ..................... | .................... | .................... | .................... |
| F | 34 | hCoV-19/USA/WA-S2626/2020\|EPI_ISL_497258\|2020-06-1 | .................... | .................... | ..................... | .................... | .................... | .................... |
| F | 35 | hCoV-19/USA/WA-S2627/2020\|EPI_ISL_497259\|2020-06-2 | .................... | .................... | ..................... | .................... | .................... | .................... |
| F | 36 | hCoV-19/USA/WA-S2628/2020\|EPI_ISL_497260\|2020-06-2 | .................... | .................... | ..................... | .................... | .................... | .................... |
| F | 37 | hCoV-19/USA/WA-S2629/2020\|EPI_ISL_497261\|2020-06-2 | .................... | .................... | ..................... | .................... | .................... | .................... |
| F | 38 | hCoV-19/USA/WA-S2630/2020\|EPI_ISL_497262\|2020-06-2 | .................... | .................... | ..................... | .................... | .................... | .................... |
| F | 39 | hCoV-19/USA/WA-S2631/2020\|EPI_ISL_497263\|2020-06-2 | .................... | .................... | ..................... | .................... | .................... | .................... |
| F | 40 | hCoV-19/USA/WA-S2633/2020\|EPI_ISL_497265\|2020-06-2 | .................... | .................... | ..................... | .................... | .................... | .................... |
| F | 41 | hCoV-19/USA/WA-S2634/2020\|EPI_ISL_497266\|2020-06-2 | .................... | .................... | ..................... | .................... | .................... | .................... |
| F | 42 | hCoV-19/USA/WA-S2635/2020\|EPI_ISL_497267\|2020-06-2 | .................... | .................... | ..................... | .................... | .................... | .................... |

```
F    43 hCoV-19/USA/WA-S2636/2020|EPI_ISL_497268|2020-06-2   ....................  ....................  ....................  ....................  ....................  ....................
F    44 hCoV-19/USA/WA-S2637/2020|EPI_ISL_497269|2020-06-2   ....................  ....................  ....................  ....................  ....................  ....................
F    45 hCoV-19/USA/WA-S2638/2020|EPI_ISL_497270|2020-06-2   ....................  ....................  ....................  ....................  ....................  ....................
F    46 hCoV-19/USA/WA-S2639/2020|EPI_ISL_497271|2020-06-2   ....................  ....................  ....................  ....................  ....................  ....................
F    47 hCoV-19/USA/WA-S2640/2020|EPI_ISL_497272|2020-06-2   ....................  ....................  ....................  ....................  ....................  ....................
F    48 hCoV-19/USA/WA-S2641/2020|EPI_ISL_497273|2020-06-2   ....................  ....................  ....................  ....................  ....................  ....................
F    49 hCoV-19/USA/WA-S2642/2020|EPI_ISL_497274|2020-06-2   ....................  ....................  ....................  ....................  ....................  ....................
F    50 hCoV-19/USA/WA-S2643/2020|EPI_ISL_497275|2020-06-2   ....................  ....................  ....................  ....................  ....................  ....................
F    51 hCoV-19/USA/WA-S2644/2020|EPI_ISL_497276|2020-06-2   ....................  ....................  ....................  ....................  ....................  ....................
F    52 hCoV-19/USA/WA-S2645/2020|EPI_ISL_497277|2020-06-2   ....................  ....................  ....................  ....................  ....................  ....................
F    53 hCoV-19/USA/WA-S2646/2020|EPI_ISL_497278|2020-06-2   ....................  ....................  ....................  ....................  ....................  ....................
F    54 hCoV-19/USA/WA-S2647/2020|EPI_ISL_497279|2020-06-2   ....................  ....................  ....................  ....................  ....................  ....................
F    55 hCoV-19/USA/WA-S2648/2020|EPI_ISL_497280|2020-06-2   ....................  ....................  ....................  ....................  ....................  ....................
F    56 hCoV-19/USA/WA-S2649/2020|EPI_ISL_497281|2020-06-2   ....................  ....................  ....................  ....................  ....................  ....................
F    57 hCoV-19/USA/WA-S2650/2020|EPI_ISL_497282|2020-06-2   ....................  ....................  ....................  ....................  ....................  ....................
F    58 hCoV-19/USA/WA-S2651/2020|EPI_ISL_497283|2020-06-2   ....................  ....................  ....................  ....................  ....................  ....................
F    59 hCoV-19/USA/WA-S2652/2020|EPI_ISL_497284|2020-06-2   ....................  ....................  ....................  ....................  ....................  ....................
F    60 hCoV-19/USA/WA-S2653/2020|EPI_ISL_497285|2020-06-2   ....................  ....................  ....................  ....................  ....................  ....................
F    61 hCoV-19/USA/WA-S2654/2020|EPI_ISL_497286|2020-06-2   ....................  ....................  ....................  ....................  ....................  ....................
F    62 hCoV-19/USA/WA-S2655/2020|EPI_ISL_497287|2020-06-2   ....................  ....................  ....................  ....................  ....................  ....................
F    63 hCoV-19/USA/WA-S2656/2020|EPI_ISL_497288|2020-06-2   ....................  ....................  ....................  ....................  ....................  ....................
F    64 hCoV-19/USA/WA-S2657/2020|EPI_ISL_497289|2020-06-2   ....................  ....................  ....................  ....................  ....................  ....................
F    65 hCoV-19/USA/WA-S2658/2020|EPI_ISL_497290|2020-06-2   ....................  ....................  ....................  ....................  ....................  ....................
F    66 hCoV-19/USA/WA-S2659/2020|EPI_ISL_497291|2020-06-2   ....................  ....................  ....................  ....................  ....................  ....................
F    67 hCoV-19/USA/WA-S2660/2020|EPI_ISL_497292|2020-06-2   ....................  ....................  ....................  ....................  ....................  ....................
F    68 hCoV-19/USA/WA-S2661/2020|EPI_ISL_497293|2020-06-2   ....................  ....................  ....................  ....................  ....................  ....................
F    69 hCoV-19/USA/WA-S2662/2020|EPI_ISL_497294|2020-06-2   ....................  ....................  ....................  ....................  ....................  ....................
F    70 hCoV-19/USA/WA-S2663/2020|EPI_ISL_497295|2020-06-2   ....................  ....................  ....................  ....................  ....................  ....................
F    71 hCoV-19/USA/WA-S2664/2020|EPI_ISL_497296|2020-06-2   ....................  ....................  ....................  ....................  ....................  ....................
F    72 hCoV-19/USA/WA-S2665/2020|EPI_ISL_497297|2020-06-2   ....................  ....................  ....................  ....................  ....................  ....................
F    73 hCoV-19/USA/WA-S2666/2020|EPI_ISL_497298|2020-06-2   ....................  ....................  ....................  ....................  ....................  ....................
F    74 hCoV-19/USA/WA-S2667/2020|EPI_ISL_497299|2020-06-1   ....................  ....................  ....................  ....................  ....................  ....................
F    75 hCoV-19/Australia/ACT0077/2020|EPI_ISL_498537|2020   ....................  ....................  ....................  ....................  ....................  ....................
F    76 hCoV-19/Australia/ACT0082/2020|EPI_ISL_498542|2020   ....................  ....................  ....................  ....................  ....................  ....................
F    77 hCoV-19/Australia/ACT0084/2020|EPI_ISL_498543|2020   ....................  ....................  ....................  ....................  ....................  ....................
F    78 hCoV-19/Australia/ACT0086/2020|EPI_ISL_498544|2020   ....................  ....................  ....................  ....................  ....................  ....................
F    79 hCoV-19/Australia/ACT0087/2020|EPI_ISL_498545|2020   ....................  ....................  ....................  ....................  ....................  ....................
F    80 hCoV-19/Australia/ACT0088/2020|EPI_ISL_498546|2020   ....................  ....................  ....................  ....................  ....................  ....................
F    81 hCoV-19/Australia/ACT0089/2020|EPI_ISL_498547|2020   ....................  ....................  ....................  ....................  ....................  ....................
F    82 hCoV-19/Australia/ACT0090/2020|EPI_ISL_498548|2020   ....................  ....................  ....................  ....................  ....................  ....................
F    83 hCoV-19/Bangladesh/BCSIR-NILMRC-290/2020|EPI_ISL_4   ....................  ....................  ....................  ....................  ....................  ....................
F    84 hCoV-19/Bangladesh/BCSIR-NILMRC-291/2020|EPI_ISL_4   ....................  ....................  ....................  ....................  ....................  ....................
F    85 hCoV-19/Lebanon/LB-R8/2020|EPI_ISL_498551|2020-03-   ....................  ....................  ....................  ....................  ....................  ....................
F    86 hCoV-19/Lebanon/LB-LR9/2020|EPI_ISL_498552|2020-03   ....................  ....................  ....................  ....................  ....................  ....................
F    87 hCoV-19/Lebanon/LB-LR10/2020|EPI_ISL_498554|2020-0   ....................  ....................  ....................  ....................  ....................  ....................
F    88 hCoV-19/Singapore/683/2020|EPI_ISL_498581|2020-07-   ....................  ....................  ....................  ....................  ....................  ....................
F    89 hCoV-19/Singapore/684/2020|EPI_ISL_498582|2020-07-   ....................  ....................  ....................  ....................  ....................  ....................
F    90 hCoV-19/Singapore/685/2020|EPI_ISL_498583|2020-07-   ....................  ....................  ....................  ....................  ....................  ....................
F    91 hCoV-19/Singapore/686/2020|EPI_ISL_498584|2020-07-   ....................  ....................  ....................  ....................  ....................  ....................
F    92 hCoV-19/Singapore/687/2020|EPI_ISL_498585|2020-07-   ....................  ....................  ....................  ....................  ....................  ....................
F    93 hCoV-19/Singapore/688/2020|EPI_ISL_498586|2020-07-   ....................  .A..................  ....................  ....................  ....................  ..A.................
F    94 hCoV-19/Singapore/689/2020|EPI_ISL_498587|2020-07-   ....................  ....................  ....................  ....................  ....................  ....................
F    95 hCoV-19/Singapore/690/2020|EPI_ISL_498588|2020-07-   ....................  ....................  ....................  ....................  ....................  ....................
F    96 hCoV-19/Singapore/691/2020|EPI_ISL_498589|2020-07-   ....................  ....................  ....................  ....................  ....................  ..R.................
F    97 hCoV-19/Singapore/692/2020|EPI_ISL_498590|2020-07-   ....................  ....................  ....................  ....................  ....................  ....................
F    98 hCoV-19/Singapore/693/2020|EPI_ISL_498591|2020-07-   ....................  ....................  ....................  ....................  ....................  ....................
F    99 hCoV-19/Singapore/694/2020|EPI_ISL_498592|2020-07-   ....................  ....................  ....................  ....................  ....................  ....................
F   100 hCoV-19/Singapore/695/2020|EPI_ISL_498593|2020-07-   ....................  ....................  ....................  ....................  ....................  ....................
F   101 hCoV-19/Singapore/696/2020|EPI_ISL_498594|2020-07-   ....................  ....................  ....................  ....................  ....................  ....................
F   102 hCoV-19/Singapore/697/2020|EPI_ISL_498595|2020-07-   ....................  ....................  ....................  ....................  ....................  ....................
F   103 hCoV-19/Northern Ireland/NIRE-106BCC/2020|EPI_ISL_   ....................  ....................  ....................  ....................  ....................  ....................
F   104 hCoV-19/Northern Ireland/NIRE-1064EF/2020|EPI_ISL_   ....................  ....................  ....................  ....................  ....................  ....................
F   105 hCoV-19/Northern Ireland/NIRE-106B08/2020|EPI_ISL_   ....................  ....................  ....................  ....................  ....................  ....................
F   106 hCoV-19/Northern Ireland/NIRE-106B17/2020|EPI_ISL_   ....................  ....................  ....................  ....................  ....................  ....................
F   107 hCoV-19/Northern Ireland/NIRE-106740/2020|EPI_ISL_   ....................  ....................  ....................  ....................  ....................  ....................
F   108 hCoV-19/Northern Ireland/NIRE-106731/2020|EPI_ISL_   ....................  ....................  ....................  ....................  ....................  ....................
F   109 hCoV-19/Northern Ireland/NIRE-1066CB/2020|EPI_ISL_   ....................  ....................  ....................  ....................  ....................  ....................
F   110 hCoV-19/Northern Ireland/NIRE-106EF0/2020|EPI_ISL_   ....................  ....................  ....................  ....................  ....................  ....................
F   111 hCoV-19/Northern Ireland/NIRE-106898/2020|EPI_ISL_   ....................  ....................  ....................  ....................  ....................  ....................
F   112 hCoV-19/Northern Ireland/NIRE-1065FB/2020|EPI_ISL_   ....................  ....................  ....................  ....................  ....................  ....................
F   113 hCoV-19/Northern Ireland/NIRE-106625/2020|EPI_ISL_   ....................  ....................  ....................  ....................  ....................  ....................
F   114 hCoV-19/Northern Ireland/NIRE-106EC3/2020|EPI_ISL_   ....................  ....................  ....................  ....................  ....................  ....................
F   115 hCoV-19/Northern Ireland/NIRE-1064A3/2020|EPI_ISL_   ....................  ....................  ....................  ....................  ....................  ....................
F   116 hCoV-19/Northern Ireland/NIRE-10676E/2020|EPI_ISL_   ....................  ....................  ....................  ....................  ....................  ....................
F   117 hCoV-19/Northern Ireland/NIRE-106573/2020|EPI_ISL_   ....................  ....................  ....................  ....................  ....................  ....................
F   118 hCoV-19/England/NORT-29B9B2/2020|EPI_ISL_507120|20   ....................  ....................  ....................  ....................  ....................  ....................
F   119 hCoV-19/England/NORT-29BA0A/2020|EPI_ISL_507121|20   ....................  ....................  ....................  ....................  ....................  ....................
F   120 hCoV-19/England/NORT-29BE3B/2020|EPI_ISL_507123|20   ....................  ....................  ....................  ....................  ....................  ....................
F   121 hCoV-19/England/NORT-29BE4A/2020|EPI_ISL_507124|20   ....................  ....................  ....................  ....................  ....................  ....................
F   122 hCoV-19/England/NORT-29BF65/2020|EPI_ISL_507125|20   ....................  ....................  ....................  ....................  ....................  ....................
```

| | | | | | | | |
|---|---|---|---|---|---|---|---|
| F | 123 hCoV-19/England/NORT-2A61E9/2020\|EPI_ISL_507126\|20 | ···················· | ···················· | ···················· | ···················· | ···················· | ···················· |
| F | 124 hCoV-19/England/NORT-2A6213/2020\|EPI_ISL_507127\|20 | ···················· | ···················· | ···················· | ···················· | ···················· | ···················· |
| F | 125 hCoV-19/England/NORT-2A62E6/2020\|EPI_ISL_507129\|20 | ···················· | ···················· | ···················· | ···················· | ···················· | ···················· |
| F | 126 hCoV-19/England/PORT-2D1BBA/2020\|EPI_ISL_507131\|20 | ···················· | ···················· | ···················· | ···················· | ···················· | ···················· |
| F | 127 hCoV-19/England/PORT-2D1BC9/2020\|EPI_ISL_507132\|20 | ···················· | ···················· | ···················· | ···················· | ···················· | ···················· |
| F | 128 hCoV-19/England/PORT-2D1BD8/2020\|EPI_ISL_507133\|20 | ···················· | ···················· | ···················· | ···················· | ···················· | ···················· |
| F | 129 hCoV-19/England/PORT-2D1BE7/2020\|EPI_ISL_507134\|20 | ···················· | ···················· | ···················· | ···················· | ···················· | ···················· |
| F | 130 hCoV-19/England/PORT-2D1BF6/2020\|EPI_ISL_507135\|20 | ···················· | ···················· | ···················· | ···················· | ···················· | ···················· |
| F | 131 hCoV-19/England/PORT-2D225A/2020\|EPI_ISL_507137\|20 | ···················· | ···················· | ···················· | ···················· | ···················· | ···················· |
| F | 132 hCoV-19/England/PORT-2D2296/2020\|EPI_ISL_507139\|20 | ···················· | ···T················ | ···················· | ···················· | ···················· | ···················· |
| F | 133 hCoV-19/England/PORT-2D22A5/2020\|EPI_ISL_507140\|20 | ···················· | ···················· | ···················· | ···················· | ···················· | ···················· |
| F | 134 hCoV-19/England/PORT-2D23FD/2020\|EPI_ISL_507141\|20 | ···················· | ···T················ | ···················· | ···················· | ···················· | ···················· |
| F | 135 hCoV-19/England/PORT-2D2418/2020\|EPI_ISL_507142\|20 | ···················· | ···················· | ···················· | ···················· | ···················· | ···················· |
| F | 136 hCoV-19/England/PORT-2D2445/2020\|EPI_ISL_507143\|20 | ···················· | ···················· | ···················· | ···················· | ···················· | ···················· |
| F | 137 hCoV-19/England/PORT-2D2481/2020\|EPI_ISL_507146\|20 | ···················· | ···················· | ···················· | ···················· | ···················· | ···················· |
| F | 138 hCoV-19/England/PORT-2D2490/2020\|EPI_ISL_507147\|20 | ···················· | ···················· | ···················· | ···················· | ···················· | ···················· |
| F | 139 hCoV-19/England/PORT-2D24AF/2020\|EPI_ISL_507148\|20 | ···················· | ···················· | ···················· | ···················· | ···················· | ···················· |
| F | 140 hCoV-19/England/PORT-2D24BE/2020\|EPI_ISL_507149\|20 | ···················· | ···················· | ···················· | ···················· | ···················· | ···················· |
| F | 141 hCoV-19/England/PORT-2D2506/2020\|EPI_ISL_507152\|20 | ···················· | ···················· | ···················· | ···················· | ···················· | ···················· |
| F | 142 hCoV-19/England/PORT-2D2515/2020\|EPI_ISL_507153\|20 | ···················· | ···················· | ···················· | ···················· | ···················· | ···················· |
| F | 143 hCoV-19/England/PORT-2D2533/2020\|EPI_ISL_507155\|20 | ···················· | ···················· | ···················· | ···················· | ···················· | ···················· |
| F | 144 hCoV-19/England/PORT-2D4003/2020\|EPI_ISL_507159\|20 | ···················· | ···T················ | ···················· | ···················· | ···················· | ···················· |
| F | 145 hCoV-19/England/SHEF-C2389/2020\|EPI_ISL_507161\|202 | ···················· | ···················· | ···················· | ···················· | ···················· | ···················· |
| F | 146 hCoV-19/England/SHEF-C433C/2020\|EPI_ISL_507163\|202 | ···················· | ···················· | ···················· | ···················· | ···················· | ···················· |
| F | 147 hCoV-19/England/SHEF-C5B51/2020\|EPI_ISL_507165\|202 | ···················· | ···················· | ···················· | ···················· | ···················· | ···················· |
| F | 148 hCoV-19/England/SHEF-D32E2/2020\|EPI_ISL_507178\|202 | ···················· | ···················· | ···················· | ···················· | ···················· | ···················· |
| F | 149 hCoV-19/England/SHEF-D34BF/2020\|EPI_ISL_507179\|202 | ···················· | ···················· | ···················· | ···················· | ···················· | ···················· |
| F | 150 hCoV-19/Scotland/EDB1334/2020\|EPI_ISL_507180\|2020- | ···················· | ·A·················· | ···················· | ···················· | ···················· | ···················· |
| F | 151 hCoV-19/Scotland/EDB3854/2020\|EPI_ISL_507181\|2020- | ···················· | ·A·················· | ···················· | ···················· | ···················· | ···················· |
| F | 152 hCoV-19/Scotland/EDB6683/2020\|EPI_ISL_507184\|2020- | ···················· | ···················· | ···················· | ···················· | ···················· | ···················· |
| F | 153 hCoV-19/Scotland/EDB6684/2020\|EPI_ISL_507185\|2020- | ···················· | ···················· | ···················· | ···················· | ···················· | ···················· |
| F | 154 hCoV-19/Scotland/EDB6693/2020\|EPI_ISL_507188\|2020- | ···················· | ···················· | ···················· | ···················· | ···················· | ···················· |
| F | 155 hCoV-19/Scotland/EDB6703/2020\|EPI_ISL_507192\|2020- | ···················· | ···················· | ···················· | ···················· | ···················· | ···················· |
| F | 156 hCoV-19/Scotland/EDB6710/2020\|EPI_ISL_507196\|2020- | ···················· | ·A·················· | ···················· | ···················· | ···················· | ···················· |
| F | 157 hCoV-19/India/WB-S47/2020\|EPI_ISL_508450\|2020-05-0 | ···················· | ···················· | ···················· | ···················· | ···················· | ···················· |
| F | 158 hCoV-19/India/WB-S54/2020\|EPI_ISL_508451\|2020-05-0 | ···················· | ···················· | ···················· | ···················· | ···················· | ···················· |
| F | 159 hCoV-19/India/WB-S57/2020\|EPI_ISL_508452\|2020-05-0 | ···················· | ···················· | ···················· | ···················· | ···················· | ···················· |
| F | 160 hCoV-19/India/WB-S63/2020\|EPI_ISL_508453\|2020-05-2 | ···················· | ···················· | ···················· | ···················· | ···················· | ···················· |
| F | 161 hCoV-19/India/WB-S64/2020\|EPI_ISL_508454\|2020-05-2 | ···················· | ·A·················· | ···················· | ···················· | ···················· | ··A················· |
| F | 162 hCoV-19/India/WB-S66/2020\|EPI_ISL_508456\|2020-05-2 | ···················· | ···················· | ···················· | ···················· | ···················· | ···················· |
| F | 163 hCoV-19/India/WB-S67/2020\|EPI_ISL_508457\|2020-05-2 | ···················· | ···················· | ···················· | ···················· | ···················· | ···················· |
| F | 164 hCoV-19/India/WB-S70/2020\|EPI_ISL_508460\|2020-05-2 | ···················· | ···················· | ···················· | ···················· | ···················· | ···················· |
| F | 165 hCoV-19/India/WB-S73/2020\|EPI_ISL_508463\|2020-05-2 | ···················· | ···················· | ···················· | ···················· | ···················· | ···················· |
| F | 166 hCoV-19/India/WB-S74/2020\|EPI_ISL_508464\|2020-05-2 | ···················· | ···················· | ···················· | ···················· | ···················· | ···················· |
| F | 167 hCoV-19/India/WB-S75/2020\|EPI_ISL_508465\|2020-05-2 | ···················· | ···················· | ···················· | ···················· | ···················· | ···················· |
| F | 168 hCoV-19/India/WB-S78/2020\|EPI_ISL_508468\|2020-05-2 | ···················· | ···················· | ···················· | ···················· | ···················· | ···················· |
| F | 169 hCoV-19/India/WB-S79/2020\|EPI_ISL_508469\|2020-05-2 | ···················· | ···················· | ···················· | ···················· | ···················· | ···················· |
| F | 170 hCoV-19/India/WB-S81/2020\|EPI_ISL_508471\|2020-05-2 | ···················· | ···················· | ···················· | ···················· | ···················· | ···················· |
| F | 171 hCoV-19/India/WB-S86/2020\|EPI_ISL_508475\|2020-05-2 | ···················· | ···················· | ···················· | ···················· | ···················· | ···················· |
| F | 172 hCoV-19/India/WB-S92/2020\|EPI_ISL_508481\|2020-05-2 | ···················· | ···················· | ···················· | ···················· | ···················· | ···················· |
| F | 173 hCoV-19/India/WB-S93/2020\|EPI_ISL_508482\|2020-06-0 | ···················· | ···················· | ···················· | ···················· | ···················· | ···················· |
| F | 174 hCoV-19/India/WB-S94/2020\|EPI_ISL_508483\|2020-06-0 | ···················· | ···················· | ···················· | ···················· | ···················· | ···················· |
| F | 175 hCoV-19/India/WB-S98/2020\|EPI_ISL_508487\|2020-06-1 | ···················· | ···················· | ···················· | ···················· | ···················· | ···················· |
| F | 176 hCoV-19/India/HR-TF1/2020\|EPI_ISL_508489\|2020-06-1 | ···················· | ·A·················· | ···················· | ···················· | ···················· | ··A················· |
| F | 177 hCoV-19/India/HR-TF10/2020\|EPI_ISL_508490\|2020-06- | ···················· | ···················· | ···················· | ···················· | ···················· | ···················· |
| F | 178 hCoV-19/India/HR-TF12/2020\|EPI_ISL_508491\|2020-06- | ···················· | ·A·················· | ···················· | ···················· | ···················· | ··A················· |
| F | 179 hCoV-19/India/HR-TF13/2020\|EPI_ISL_508492\|2020-06- | ···················· | ·A·················· | ···················· | ···················· | ···················· | ··A················· |
| F | 180 hCoV-19/India/HR-TF14/2020\|EPI_ISL_508493\|2020-06- | ···················· | ·A·················· | ···················· | ···················· | ···················· | ··A················· |
| F | 181 hCoV-19/India/HR-TF15/2020\|EPI_ISL_508494\|2020-06- | ···················· | ·A·················· | ···················· | ···················· | ···················· | ··A················· |
| F | 182 hCoV-19/India/DL-TF17/2020\|EPI_ISL_508495\|2020-06- | ···················· | ·A·················· | ···················· | ···················· | ···················· | ··A················· |
| F | 183 hCoV-19/India/HR-TF2/2020\|EPI_ISL_508496\|2020-06-1 | ···················· | ·A·················· | ···················· | ···················· | ···················· | ··A················· |
| F | 184 hCoV-19/India/HR-TF20/2020\|EPI_ISL_508497\|2020-06- | ···················· | ·A·················· | ···················· | ···················· | ···················· | ··A················· |
| F | 185 hCoV-19/India/HR-TF24/2020\|EPI_ISL_508498\|2020-06- | ···················· | ···················· | ···················· | ···················· | ···················· | ···················· |
| F | 186 hCoV-19/India/HR-TF25/2020\|EPI_ISL_508499\|2020-06- | ···················· | ·A·················· | ···················· | ···················· | ···················· | ··A················· |
| F | 187 hCoV-19/USA/FL-BPHL-0722/2020\|EPI_ISL_509780\|2020- | ···················· | ···················· | ···················· | ···················· | ···················· | ···················· |
| F | 188 hCoV-19/USA/FL-BPHL-0723/2020\|EPI_ISL_509781\|2020- | ···················· | ···················· | ···················· | ···················· | ···················· | ···················· |
| F | 190 hCoV-19/USA/FL-BPHL-0725/2020\|EPI_ISL_509783\|2020- | ···················· | ···················· | ···················· | ···················· | ···················· | ···················· |
| F | 191 hCoV-19/USA/FL-BPHL-0728/2020\|EPI_ISL_509786\|2020- | ···················· | ···················· | ···················· | ···················· | ···················· | ···················· |
| F | 192 hCoV-19/USA/FL-BPHL-0729/2020\|EPI_ISL_509787\|2020- | ···················· | ···················· | ···················· | ···················· | ···················· | ···················· |
| F | 193 hCoV-19/USA/FL-BPHL-0730/2020\|EPI_ISL_509788\|2020- | ···················· | ···················· | ···················· | ···················· | ···················· | ···················· |
| F | 194 hCoV-19/USA/FL-BPHL-0731/2020\|EPI_ISL_509789\|2020- | ···················· | ···················· | ···················· | ···················· | ···················· | ···················· |
| F | 195 hCoV-19/USA/FL-BPHL-0733/2020\|EPI_ISL_509791\|2020- | ···················· | ···················· | ···················· | ···················· | ···················· | ···················· |
| F | 196 hCoV-19/USA/FL-BPHL-0734/2020\|EPI_ISL_509792\|2020- | ···················· | ···················· | ···················· | ···················· | ···················· | ···················· |
| F | 197 hCoV-19/USA/FL-BPHL-0735/2020\|EPI_ISL_509793\|2020- | ···················· | ···················· | ···················· | ···················· | ···················· | ···················· |
| F | 198 hCoV-19/USA/FL-BPHL-0736/2020\|EPI_ISL_509794\|2020- | ···················· | ···················· | ···················· | ···················· | ···················· | ···················· |
| F | 199 hCoV-19/USA/FL-BPHL-0737/2020\|EPI_ISL_509795\|2020- | ···················· | ···················· | ···················· | ···················· | ···················· | ···················· |
| F | 201 hCoV-19/USA/FL-BPHL-0739/2020\|EPI_ISL_509797\|2020- | ···················· | ···················· | ···················· | ···················· | ···················· | ···················· |
| F | 202 hCoV-19/USA/CA-UW-869/2020\|EPI_ISL_509799\|2020-07- | ···················· | ···················· | ···················· | ···················· | ···················· | ···················· |
| F | 203 hCoV-19/Switzerland/BL-ETHZ-210019/2020\|EPI_ISL_51 | ···················· | ···················· | ···················· | ···················· | ···················· | ···················· |
| F | 204 hCoV-19/Switzerland/ZH-ETHZ-210021/2020\|EPI_ISL_51 | ···················· | ···················· | ···················· | ···················· | ···················· | ···················· |

```
F 205 hCoV-19/Switzerland/ZH-ETHZ-210022/2020|EPI_ISL_51 ···················· ···················· ···················· ···················· ···················· ····················
F 206 hCoV-19/Switzerland/ZH-ETHZ-210023/2020|EPI_ISL_51 ···················· ···················· ···················· ···················· ···················· ····················
F 207 hCoV-19/Switzerland/TI-ETHZ-210025/2020|EPI_ISL_51 ···················· ···················· ···················· ···················· ···················· ····················
F 208 hCoV-19/Switzerland/UR-ETHZ-210026/2020|EPI_ISL_51 ···················· ···················· ···················· ···················· ·······S············ ····················
F 209 hCoV-19/Switzerland/AG-ETHZ-210028/2020|EPI_ISL_51 ···················· ···················· ···················· ···················· ···················· ····················
F 210 hCoV-19/Switzerland/ZH-ETHZ-210030/2020|EPI_ISL_51 ···················· ···················· ···················· ···················· ···················· ····················
F 211 hCoV-19/Switzerland/ZH-ETHZ-210032/2020|EPI_ISL_51 ···················· ···················· ···················· ···················· ···················· ····················
F 212 hCoV-19/Switzerland/ZH-ETHZ-210033/2020|EPI_ISL_51 ···················· ···················· ···················· ···················· ···················· ····················
F 213 hCoV-19/Switzerland/ZH-ETHZ-210034/2020|EPI_ISL_51 ···················· ···················· ···················· ···················· ···················· ····················
F 214 hCoV-19/Switzerland/BE-ETHZ-210035/2020|EPI_ISL_51 ···········K········ ···················· ···················· ···················· ···················· ····················
F 215 hCoV-19/Switzerland/GR-ETHZ-210036/2020|EPI_ISL_51 ···········T········ ···················· ···················· ···················· ···················· ····················
F 216 hCoV-19/Switzerland/ZH-ETHZ-210037/2020|EPI_ISL_51 ···················· ···················· ···················· ···················· ···················· ····················
F 217 hCoV-19/Switzerland/BE-ETHZ-210038/2020|EPI_ISL_51 ···················· ···················· ···················· ···················· ···················· ····················
F 218 hCoV-19/Switzerland/BE-ETHZ-210039/2020|EPI_ISL_51 ···················· ···················· ···················· ···················· ···················· ····················
F 219 hCoV-19/Switzerland/SZ-ETHZ-210040/2020|EPI_ISL_51 ···················· ···················· ···················· ···················· ···················· ····················
F 220 hCoV-19/Switzerland/GR-ETHZ-210045/2020|EPI_ISL_51 ···········T········ ···················· ···················· ···················· ···················· ····················
F 221 hCoV-19/Switzerland/ZH-ETHZ-210047/2020|EPI_ISL_51 ···················· ···················· ···················· ···················· ···················· ····················
F 222 hCoV-19/Switzerland/SZ-ETHZ-210048/2020|EPI_ISL_51 ···················· ···················· ···················· ···················· ···················· ····················
F 223 hCoV-19/Switzerland/BL-ETHZ-210050/2020|EPI_ISL_51 ···················· ···················· ···················· ···················· ···················· ····················
F 224 hCoV-19/Switzerland/ZH-ETHZ-210051/2020|EPI_ISL_51 ···················· ···················· ···················· ···················· ·······S············ ······R·············
F 225 hCoV-19/Switzerland/UR-ETHZ-210055/2020|EPI_ISL_51 ···················· ·A·················· ···················· ···················· ···················· ····················
F 226 hCoV-19/Switzerland/BE-ETHZ-210060/2020|EPI_ISL_51 ···········T········ ·········R·········· ···················· ···················· ···················· ····················
F 227 hCoV-19/Switzerland/VD-ETHZ-210061/2020|EPI_ISL_51 ···················· ···················· ···················· ···················· ···················· ····················
F 228 hCoV-19/Switzerland/UR-ETHZ-210062/2020|EPI_ISL_51 ···················· ···················· ···················· ···················· ···················· ····················
F 229 hCoV-19/Switzerland/BL-ETHZ-210064/2020|EPI_ISL_51 ···················· ···················· ···················· ···················· ···················· ····················
F 230 hCoV-19/Switzerland/BE-ETHZ-210066/2020|EPI_ISL_51 ···················· ···················· ···················· ···················· ···················· ····················
F 231 hCoV-19/Israel/CVL-n14268/2020|EPI_ISL_514308|2020 ···················· ···················· ···················· ···················· ···················· ····················
F 232 hCoV-19/Israel/CVL-n14269/2020|EPI_ISL_514309|2020 ···················· ···················· ···················· ···················· ···················· ····················
F 233 hCoV-19/Israel/CVL-n14270/2020|EPI_ISL_514310|2020 ···················· ···················· ···················· ···················· ···················· ····················
F 234 hCoV-19/Israel/CVL-n14279/2020|EPI_ISL_514313|2020 ···················· ···················· ···················· ···················· ···················· ····················
F 235 hCoV-19/Israel/CVL-n14285/2020|EPI_ISL_514314|2020 ···················· ···················· ···················· ···················· ···················· ····················
F 236 hCoV-19/Israel/CVL-n14292/2020|EPI_ISL_514315|2020 ···················· ···················· ···················· ···················· ···················· ····················
F 237 hCoV-19/Israel/CVL-n14293/2020|EPI_ISL_514316|2020 ···················· ···················· ···················· ···················· ···················· ····················
F 238 hCoV-19/Peru/LOR-INS-058/2020|EPI_ISL_514317|2020- ···················· ···················· ···················· ···················· ···················· ····················
F 239 hCoV-19/South Africa/KRISP-K002253/2020|EPI_ISL_51 ···················· ···················· ···················· ···················· ···················· ····················
F 240 hCoV-19/South Africa/KRISP-K002255/2020|EPI_ISL_51 ···················· ···················· ···················· ···················· ···················· ····················
F 241 hCoV-19/South Africa/KRISP-K002256/2020|EPI_ISL_51 ···················· ···················· ···················· ···················· ···················· ····················
F 242 hCoV-19/South Africa/KRISP-K002258/2020|EPI_ISL_51 ···················· ···················· ···················· ···················· ···················· ····················
F 243 hCoV-19/South Africa/KRISP-K002259/2020|EPI_ISL_51 ···················· ···················· ···················· ···················· ···················· ····················
F 244 hCoV-19/South Africa/KRISP-K002260/2020|EPI_ISL_51 ···················· ···················· ···················· ···················· ···················· ····················
F 245 hCoV-19/South Africa/KRISP-K002264/2020|EPI_ISL_51 ···················· ···················· ···················· ···················· ···················· ····················
F 246 hCoV-19/South Africa/KRISP-K002266/2020|EPI_ISL_51 ···················· ···················· ···················· ···················· ···················· ····················
F 247 hCoV-19/South Africa/KRISP-K002268/2020|EPI_ISL_51 ···················· ···················· ···················· ···················· ···················· ····················
F 248 hCoV-19/South Africa/KRISP-K002269/2020|EPI_ISL_51 ···················· ···················· ···················· ···················· ···················· ····················
F 249 hCoV-19/South Africa/KRISP-K002270/2020|EPI_ISL_51 ···················· ···················· ···················· ···················· ···················· ····················
F 250 hCoV-19/South Africa/KRISP-K002271/2020|EPI_ISL_51 ···················· ···················· ···················· ···················· ···················· ····················
F 251 hCoV-19/South Africa/KRISP-K002273/2020|EPI_ISL_51 ···················· ···················· ···················· ···················· ···················· ····················
F 252 hCoV-19/South Africa/KRISP-K002274/2020|EPI_ISL_51 ···················· ···················· ···················· ···················· ···················· ····················
F 253 hCoV-19/South Africa/KRISP-K002276/2020|EPI_ISL_51 ···················· ···················· ···················· ···················· ···················· ····················
F 254 hCoV-19/South Africa/KRISP-K002277/2020|EPI_ISL_51 ···················· ···················· ···················· ···················· ···················· ····················
F 255 hCoV-19/South Africa/KRISP-K002278/2020|EPI_ISL_51 ···················· ···················· ···················· ···················· ···················· ····················
F 256 hCoV-19/South Africa/KRISP-K002279/2020|EPI_ISL_51 ···················· ···················· ···················· ···················· ···················· ····················
F 257 hCoV-19/USA/WA-S2568/2020|EPI_ISL_497200|2020-06-1 ···················· ···················· ···················· ···················· ···················· ····················
F 258 hCoV-19/USA/WA-S2569/2020|EPI_ISL_497201|2020-06-1 ···················· ···················· ···················· ···················· ···················· ····················
F 259 hCoV-19/USA/WA-S2592/2020|EPI_ISL_497224|2020-06-1 ···················· ···················· ···················· ···················· ···················· ····················
F 260 hCoV-19/USA/WA-S2593/2020|EPI_ISL_497225|2020-06-1 ···················· ···················· ···················· ···················· ···················· ····················
F 261 hCoV-19/USA/WA-S2594/2020|EPI_ISL_497226|2020-06-1 ···················· ···················· ···················· ···················· ···················· ····················
F 262 hCoV-19/USA/WA-S2596/2020|EPI_ISL_497228|2020-06-1 ···················· ···················· ···················· ···················· ···················· ····················
F 263 hCoV-19/USA/WA-S2598/2020|EPI_ISL_497230|2020-06-1 ···················· ···················· ···················· ···················· ···················· ····················
F 264 hCoV-19/USA/WA-S2599/2020|EPI_ISL_497231|2020-06-1 ···················· ·A·················· ···················· ···················· ···················· ····················
F 265 hCoV-19/USA/WA-S2600/2020|EPI_ISL_497232|2020-06-1 ···················· ···················· ···················· ···················· ···················· ····················
F 266 hCoV-19/USA/WA-S2601/2020|EPI_ISL_497233|2020-06-1 ···················· ···················· ···················· ···················· ···················· ····················
F 267 hCoV-19/USA/WA-S2602/2020|EPI_ISL_497234|2020-06-1 ···················· ···················· ···················· ···················· ···················· ····················
F 268 hCoV-19/USA/WA-S2603/2020|EPI_ISL_497235|2020-06-1 ···················· ···················· ···················· ···················· ···················· ····················
F 269 hCoV-19/USA/WA-S2604/2020|EPI_ISL_497236|2020-06-1 ···················· ···················· ···················· ···················· ···················· ····················
F 270 hCoV-19/USA/WA-S2605/2020|EPI_ISL_497237|2020-06-1 ···················· ···················· ···················· ···················· ···················· ····················
F 271 hCoV-19/USA/WA-S2606/2020|EPI_ISL_497238|2020-06-1 ···················· ···················· ···················· ···················· ···················· ····················
F 272 hCoV-19/USA/WA-S2608/2020|EPI_ISL_497240|2020-06-1 ···················· ···················· ···················· ···················· ···················· ····················
F 273 hCoV-19/USA/WA-S2609/2020|EPI_ISL_497241|2020-06-1 ···················· ···················· ···················· ···················· ···················· ····················
F 274 hCoV-19/USA/WA-S2610/2020|EPI_ISL_497242|2020-06-1 ···················· ···················· ···················· ···················· ···················· ····················
F 275 hCoV-19/USA/WA-S2611/2020|EPI_ISL_497243|2020-06-1 ···················· ···················· ···················· ···················· ···················· ····················
F 276 hCoV-19/USA/WA-S2612/2020|EPI_ISL_497244|2020-06-1 ···················· ···················· ···················· ···················· ···················· ····················
F 277 hCoV-19/Australia/ACT0033/2020|EPI_ISL_498500|2020 ···················· ···················· ···················· ···················· ···················· ····················
F 278 hCoV-19/Australia/ACT0034/2020|EPI_ISL_498501|2020 ···················· ·············A······ ···················· ···················· ···················· ····················
F 279 hCoV-19/Australia/ACT0035/2020|EPI_ISL_498502|2020 ···················· ···················· ···················· ···················· ···················· ····················
F 280 hCoV-19/Australia/ACT0036/2020|EPI_ISL_498503|2020 ···················· ···················· ···················· ···················· ···················· ····················
F 281 hCoV-19/Australia/ACT0038/2020|EPI_ISL_498505|2020 ···················· ···················· ···················· ···················· ···················· ····················
F 282 hCoV-19/Australia/ACT0039/2020|EPI_ISL_498506|2020 ···················· ···················· ···················· ···················· ···················· ····················
F 283 hCoV-19/Australia/ACT0043/2020|EPI_ISL_498510|2020 ···················· ···················· ···················· ···················· ···················· ····················
F 284 hCoV-19/Australia/ACT0044/2020|EPI_ISL_498511|2020 ···················· ···················· ···················· ···················· ···················· ····················
```

| | | | | | | | | |
|---|---|---|---|---|---|---|---|---|
| F | 285 | hCoV-19/Australia/ACT0045/2020\|EPI_ISL_498512\|2020 | ···················· | ···················· | ····················· | ···················· | ···················· | ···················· |
| F | 286 | hCoV-19/Australia/ACT0046/2020\|EPI_ISL_498513\|2020 | ···················· | ···················· | ····················· | ···················· | ···················· | ···················· |
| F | 287 | hCoV-19/Australia/ACT0047/2020\|EPI_ISL_498514\|2020 | ···················· | ···················· | ····················· | ···················· | ···················· | ···················· |
| F | 288 | hCoV-19/Australia/ACT0048/2020\|EPI_ISL_498515\|2020 | ···················· | ···················· | ····················· | ···················· | ···················· | ···················· |
| F | 289 | hCoV-19/Australia/ACT0051/2020\|EPI_ISL_498518\|2020 | ···················· | ···················· | ····················· | ···················· | ···················· | ···················· |
| F | 290 | hCoV-19/Australia/ACT0054/2020\|EPI_ISL_498520\|2020 | ···················· | ···················· | ····················· | ···················· | ···················· | ···················· |
| F | 291 | hCoV-19/Australia/ACT0059/2020\|EPI_ISL_498523\|2020 | ···················· | ···················· | ····················· | ···················· | ···················· | ···················· |
| F | 292 | hCoV-19/Australia/ACT0061/2020\|EPI_ISL_498525\|2020 | ···················· | ···················· | ····················· | ···················· | ···················· | ···················· |
| F | 293 | hCoV-19/Australia/ACT0064/2020\|EPI_ISL_498526\|2020 | ···················· | ···················· | ····················· | ···················· | ···················· | ···················· |
| F | 294 | hCoV-19/Australia/ACT0068/2020\|EPI_ISL_498529\|2020 | ···················· | ···················· | ····················· | ···················· | ···················· | ···················· |
| F | 295 | hCoV-19/Australia/ACT0071/2020\|EPI_ISL_498531\|2020 | ···················· | ····A··············· | ····················· | ···················· | ···················· | ···················· |
| F | 296 | hCoV-19/Australia/ACT0080/2020\|EPI_ISL_498540\|2020 | ···················· | ···················· | ····················· | ···················· | ···················· | ···················· |
| F | 297 | hCoV-19/Italy/FVG-ICGEB-S208/2020\|EPI_ISL_498558\|2 | ···················· | ···················· | ····················· | ···················· | ···················· | ···················· |
| F | 298 | hCoV-19/Italy/FVG-ICGEB-S211/2020\|EPI_ISL_498559\|2 | ···················· | ···················· | ····················· | ···················· | ···················· | ···················· |
| F | 299 | hCoV-19/Italy/FVG-ICGEB-S212/2020\|EPI_ISL_498560\|2 | ···················· | ···················· | ····················· | ···················· | ···················· | ···················· |
| F | 300 | hCoV-19/Italy/FVG-ICGEB-S214/2020\|EPI_ISL_498561\|2 | ···················· | ···················· | ····················· | ···················· | ···················· | ···················· |
| F | 301 | hCoV-19/Italy/FVG-ICGEB-S217/2020\|EPI_ISL_498562\|2 | ···················· | ···················· | ····················· | ···················· | ···················· | ···················· |
| F | 302 | hCoV-19/Italy/FVG-ICGEB-S218/2020\|EPI_ISL_498563\|2 | ···················· | ···················· | ····················· | ···················· | ···················· | ···················· |
| F | 303 | hCoV-19/Singapore/666/2020\|EPI_ISL_498564\|2020-07- | ···················· | ···················· | ····················· | ···················· | ···················· | ···················· |
| F | 304 | hCoV-19/Singapore/667/2020\|EPI_ISL_498565\|2020-07- | ···················· | ···················· | ····················· | ···················· | ···················· | ···················· |
| F | 305 | hCoV-19/Singapore/668/2020\|EPI_ISL_498566\|2020-05- | ···················· | ···················· | ····················· | ···················· | ···················· | ···················· |
| F | 306 | hCoV-19/Singapore/669/2020\|EPI_ISL_498567\|2020-07- | ···················· | ···················· | ····················· | ···················· | ···················· | ···················· |
| F | 307 | hCoV-19/Singapore/670/2020\|EPI_ISL_498568\|2020-07- | ···················· | ···················· | ····················· | ···················· | ···················· | ···················· |
| F | 308 | hCoV-19/Singapore/671/2020\|EPI_ISL_498569\|2020-06- | ···················· | ···················· | ····················· | ···················· | ···················· | ···················· |
| F | 309 | hCoV-19/Singapore/672/2020\|EPI_ISL_498570\|2020-06- | ···················· | ···················· | ····················· | ···················· | ···················· | ···················· |
| F | 310 | hCoV-19/Singapore/673/2020\|EPI_ISL_498571\|2020-07- | ···················· | ···················· | ····················· | ···················· | ···················· | ···················· |
| F | 311 | hCoV-19/England/NORT-29D288/2020\|EPI_ISL_499800\|20 | ···················· | ···················· | ····················· | ···················· | ···················· | ···················· |
| F | 312 | hCoV-19/England/NORT-29D2F1/2020\|EPI_ISL_499801\|20 | ···················· | ···················· | ····················· | ···················· | ···················· | ···················· |
| F | 313 | hCoV-19/England/NORT-29D376/2020\|EPI_ISL_499802\|20 | ···················· | ···················· | ····················· | ···················· | ···················· | ···················· |
| F | 314 | hCoV-19/England/NORT-29D437/2020\|EPI_ISL_499803\|20 | ···················· | ···················· | ····················· | ···················· | ···················· | ···················· |
| F | 315 | hCoV-19/England/NORT-29D455/2020\|EPI_ISL_499804\|20 | ···················· | ···················· | ····················· | ···················· | ···················· | ···················· |
| F | 316 | hCoV-19/England/NORT-29D464/2020\|EPI_ISL_499805\|20 | ···················· | ···················· | ····················· | ···················· | ···················· | ···················· |
| F | 317 | hCoV-19/England/NORT-29D4FB/2020\|EPI_ISL_499806\|20 | ···················· | ···················· | ····················· | ···················· | ···················· | ···················· |
| F | 318 | hCoV-19/England/NORT-2A0413/2020\|EPI_ISL_499807\|20 | ···················· | ···················· | ····················· | ···················· | ···················· | ···················· |
| F | 319 | hCoV-19/England/NOTT-112D5E/2020\|EPI_ISL_499808\|20 | ···················· | ···················· | ····················· | ···················· | ···················· | ···················· |
| F | 320 | hCoV-19/England/NOTT-112D6D/2020\|EPI_ISL_499809\|20 | ···················· | ···················· | ····················· | ···················· | ···················· | ···················· |
| F | 321 | hCoV-19/England/LCST-24D735A/2020\|EPI_ISL_499810\|2 | ···················· | ···················· | ····················· | ···················· | ···················· | ···················· |
| F | 322 | hCoV-19/England/LIVE-A29AB/2020\|EPI_ISL_499811\|202 | ···················· | ···················· | ····················· | ···················· | ···················· | ···················· |
| F | 325 | hCoV-19/England/LIVE-A4AF1/2020\|EPI_ISL_499814\|202 | ···················· | ···················· | ····················· | ···················· | ···················· | ···················· |
| F | 326 | hCoV-19/England/LIVE-A4B1C/2020\|EPI_ISL_499815\|202 | ···················· | ···················· | ····················· | ···················· | ···················· | ···················· |
| F | 327 | hCoV-19/England/LIVE-A4C46/2020\|EPI_ISL_499816\|202 | ···················· | ···················· | ····················· | ···················· | ···················· | ···················· |
| F | 330 | hCoV-19/England/LIVE-A5401/2020\|EPI_ISL_499819\|202 | ···················· | ···················· | ····················· | ···················· | ···················· | ···················· |
| F | 333 | hCoV-19/England/LIVE-A55E0/2020\|EPI_ISL_499822\|202 | ···················· | ···················· | ····················· | ···················· | ···················· | ···················· |
| F | 334 | hCoV-19/England/LIVE-A56FC/2020\|EPI_ISL_499823\|202 | ···················· | ···················· | ····················· | ···················· | ···················· | ···················· |
| F | 335 | hCoV-19/England/LIVE-A48D9/2020\|EPI_ISL_499824\|202 | ···················· | ···················· | ····················· | ···················· | ···················· | ···················· |
| F | 336 | hCoV-19/Northern Ireland/NIRE-106F75/2020\|EPI_ISL_ | ···················· | ······A············· | ····················· | ···················· | ···················· | ···················· |
| F | 337 | hCoV-19/Northern Ireland/NIRE-106EB4/2020\|EPI_ISL_ | ···················· | ···················· | ····················· | ···················· | ···················· | ···················· |
| F | 338 | hCoV-19/Northern Ireland/NIRE-106986/2020\|EPI_ISL_ | ···················· | ···················· | ····················· | ···················· | ···················· | ···················· |
| F | 339 | hCoV-19/Northern Ireland/NIRE-106889/2020\|EPI_ISL_ | ···················· | ·A·················· | ····················· | ···················· | ···················· | ···················· |
| F | 340 | hCoV-19/Northern Ireland/NIRE-10686B/2020\|EPI_ISL_ | ···················· | ···················· | ····················· | ···················· | ···················· | ···················· |
| F | 341 | hCoV-19/Northern Ireland/NIRE-106F39/2020\|EPI_ISL_ | ···················· | ···················· | ····················· | ···················· | ···················· | ···················· |
| F | 342 | hCoV-19/Northern Ireland/NIRE-106D11/2020\|EPI_ISL_ | ···················· | ···················· | ····················· | ···················· | ···················· | ···················· |
| F | 343 | hCoV-19/Northern Ireland/NIRE-1069C2/2020\|EPI_ISL_ | ···················· | ···················· | ····················· | ···················· | ···················· | ···················· |
| F | 344 | hCoV-19/Northern Ireland/NIRE-106AA1/2020\|EPI_ISL_ | ···················· | ···················· | ····················· | ···················· | ···················· | ···················· |
| F | 345 | hCoV-19/Northern Ireland/NIRE-106995/2020\|EPI_ISL_ | ···················· | ···················· | ····················· | ···················· | ···················· | ···················· |
| F | 346 | hCoV-19/Northern Ireland/NIRE-106564/2020\|EPI_ISL_ | ···················· | ···················· | ····················· | ···················· | ···················· | ···················· |
| F | 347 | hCoV-19/Northern Ireland/NIRE-106397/2020\|EPI_ISL_ | ···················· | ···················· | ····················· | ···················· | ···················· | ···················· |
| F | 348 | hCoV-19/Northern Ireland/NIRE-106661/2020\|EPI_ISL_ | ···················· | ···················· | ····················· | ···················· | ···················· | ···················· |
| F | 349 | hCoV-19/Northern Ireland/NIRE-106F2A/2020\|EPI_ISL_ | ···················· | ···················· | ····················· | ···················· | ···················· | ···················· |
| F | 350 | hCoV-19/Northern Ireland/NIRE-10675F/2020\|EPI_ISL_ | ···················· | ···················· | ····················· | ···················· | ···················· | ···················· |
| F | 351 | hCoV-19/Northern Ireland/NIRE-10685C/2020\|EPI_ISL_ | ···················· | ···················· | ····················· | ···················· | ···················· | ···················· |
| F | 352 | hCoV-19/Northern Ireland/NIRE-106DE4/2020\|EPI_ISL_ | ···················· | ···················· | ····················· | ···················· | ···················· | ···················· |
| F | 353 | hCoV-19/Northern Ireland/NIRE-106722/2020\|EPI_ISL_ | ···················· | ···················· | ····················· | ···················· | ···················· | ···················· |
| F | 354 | hCoV-19/Northern Ireland/NIRE-106C8D/2020\|EPI_ISL_ | ···················· | ···················· | ····················· | ···················· | ···················· | ···················· |
| F | 355 | hCoV-19/Northern Ireland/NIRE-106F48/2020\|EPI_ISL_ | ···················· | ···················· | ····················· | ···················· | ···················· | ···················· |
| F | 356 | hCoV-19/Northern Ireland/NIRE-1068A7/2020\|EPI_ISL_ | ···················· | ···················· | ····················· | ···················· | ···················· | ···················· |
| F | 357 | hCoV-19/Northern Ireland/NIRE-106B35/2020\|EPI_ISL_ | ···················· | ···················· | ····················· | ···················· | ···················· | ···················· |
| F | 358 | hCoV-19/Northern Ireland/NIRE-106670/2020\|EPI_ISL_ | ···················· | ···················· | ····················· | ···················· | ···················· | ···················· |
| F | 359 | hCoV-19/Northern Ireland/NIRE-106C23/2020\|EPI_ISL_ | ···················· | ···················· | ····················· | ···················· | ···················· | ···················· |
| F | 360 | hCoV-19/Northern Ireland/NIRE-106CF6/2020\|EPI_ISL_ | ···················· | ···················· | ····················· | ···················· | ···················· | ···················· |
| F | 361 | hCoV-19/Northern Ireland/NIRE-106D02/2020\|EPI_ISL_ | ···················· | ···················· | ····················· | ···················· | ···················· | ···················· |
| F | 362 | hCoV-19/Northern Ireland/NIRE-10678C/2020\|EPI_ISL_ | ···················· | ···················· | ····················· | ···················· | ···················· | ···················· |
| F | 363 | hCoV-19/Northern Ireland/NIRE-106B80/2020\|EPI_ISL_ | ···················· | ···················· | ····················· | ···················· | ···················· | ···················· |
| F | 364 | hCoV-19/Northern Ireland/NIRE-106CD8/2020\|EPI_ISL_ | ···················· | ···················· | ····················· | ···················· | ···················· | ···················· |
| F | 365 | hCoV-19/Northern Ireland/NIRE-1063F1/2020\|EPI_ISL_ | ···················· | ···················· | ····················· | ···················· | ···················· | ···················· |
| F | 366 | hCoV-19/Northern Ireland/NIRE-1063D3/2020\|EPI_ISL_ | ···················· | ···················· | ····················· | ···················· | ···················· | ···················· |
| F | 367 | hCoV-19/USA/AL-UW-MAYO13/2020\|EPI_ISL_501072\|2020- | ···················· | ···················· | ····················· | ···················· | ···················· | ···················· |
| F | 368 | hCoV-19/USA/CO-UW-MAYO36/2020\|EPI_ISL_501073\|2020- | ···················· | ···················· | ····················· | ···················· | ···················· | ···················· |
| F | 369 | hCoV-19/USA/GA-UW-MAYO63/2020\|EPI_ISL_501074\|2020- | ···················· | ···················· | ····················· | ···················· | ···················· | ···················· |
| F | 370 | hCoV-19/USA/IL-UW-MAYO2/2020\|EPI_ISL_501075\|2020-0 | ···················· | ···················· | ····················· | ···················· | ···················· | ···················· |

| | | | | | | | | |
|---|---|---|---|---|---|---|---|---|
| F | 371 | hCoV-19/USA/IL-UW-MAYO4/2020\|EPI_ISL_501076\|2020-0 | ···················· | ···················· | ···················· | ···················· | ···················· | ···················· |
| F | 372 | hCoV-19/USA/IL-UW-MAYO5/2020\|EPI_ISL_501077\|2020-0 | ···················· | ···················· | ···················· | ···················· | ···················· | ···················· |
| F | 373 | hCoV-19/USA/LA-UW-MAYO90/2020\|EPI_ISL_501078\|2020- | ···················· | ···················· | ···················· | ···················· | ···················· | ···················· |
| F | 374 | hCoV-19/USA/MD-UW-MAYO67/2020\|EPI_ISL_501079\|2020- | ···················· | ···················· | ···················· | ···················· | ···················· | ···················· |
| F | 375 | hCoV-19/USA/MN-UW-MAYO330/2020\|EPI_ISL_501080\|2020 | ···················· | ···················· | ···················· | ···················· | ···················· | ···················· |
| F | 376 | hCoV-19/USA/MS-UW-MAYO11/2020\|EPI_ISL_501081\|2020- | ···················· | ···················· | ···················· | ···················· | ···················· | ···················· |
| F | 377 | hCoV-19/USA/NY-UW-MAYO29/2020\|EPI_ISL_501082\|2020- | ···················· | ···················· | ···················· | ···················· | ···················· | ···················· |
| F | 378 | hCoV-19/USA/OR-UW-13394/2020\|EPI_ISL_501083\|2020-0 | ···················· | ···················· | ···················· | ···················· | ···················· | ···················· |
| F | 379 | hCoV-19/USA/UN-UW-13432/2020\|EPI_ISL_501084\|2020-0 | ···················· | ···················· | ···················· | ···················· | ···················· | ···················· |
| F | 380 | hCoV-19/USA/UN-UW-13444/2020\|EPI_ISL_501085\|2020-0 | ···················· | ···················· | ···················· | ···················· | ···················· | ···················· |
| F | 381 | hCoV-19/USA/WA-UW-10161/2020\|EPI_ISL_501086\|2020-0 | ···················· | ···················· | ···················· | ···················· | ···················· | ···················· |
| F | 382 | hCoV-19/USA/WA-UW-10212/2020\|EPI_ISL_501087\|2020-0 | ···················· | ···················· | ···················· | ···················· | ···················· | ···················· |
| F | 383 | hCoV-19/USA/WA-UW-10381/2020\|EPI_ISL_501088\|2020-0 | ···················· | ···················· | ···················· | ···················· | ···················· | ···················· |
| F | 384 | hCoV-19/USA/WA-UW-10420/2020\|EPI_ISL_501089\|2020-0 | ···················· | ···················· | ···················· | ···················· | ···················· | ···················· |
| F | 385 | hCoV-19/USA/WA-UW-10582/2020\|EPI_ISL_501090\|2020-0 | ···················· | ···················· | ···················· | ···················· | ···················· | ···················· |
| F | 386 | hCoV-19/USA/WA-UW-10613/2020\|EPI_ISL_501091\|2020-0 | ···················· | ···················· | ···················· | ···················· | ···················· | ···················· |
| F | 387 | hCoV-19/USA/WA-UW-11089/2020\|EPI_ISL_501092\|2020-0 | ···················· | ···················· | ···················· | ···················· | ···················· | ···················· |
| F | 388 | hCoV-19/USA/WA-UW-11142/2020\|EPI_ISL_501093\|2020-0 | ···················· | ···················· | ···················· | ···················· | ···················· | ···················· |
| F | 389 | hCoV-19/USA/WA-UW-11279/2020\|EPI_ISL_501095\|2020-0 | ···················· | ···················· | ···················· | ···················· | ···················· | ···················· |
| F | 390 | hCoV-19/USA/WA-UW-11293/2020\|EPI_ISL_501096\|2020-0 | ···················· | ·A·················· | ···················· | ···················· | ···················· | ···················· |
| F | 391 | hCoV-19/USA/WA-UW-11300/2020\|EPI_ISL_501097\|2020-0 | ···················· | ···················· | ···················· | ···················· | ···················· | ···················· |
| F | 392 | hCoV-19/USA/WA-UW-11399/2020\|EPI_ISL_501098\|2020-0 | ···················· | ···················· | ···················· | ···················· | ···················· | ···················· |
| F | 393 | hCoV-19/USA/WA-UW-11455/2020\|EPI_ISL_501099\|2020-0 | ···················· | ···················· | ···················· | ···················· | ···················· | ···················· |
| F | 394 | hCoV-19/USA/IL-UW-857/2020\|EPI_ISL_509800\|2020-07- | ···················· | ···················· | ···················· | ···················· | ···················· | ···················· |
| F | 395 | hCoV-19/USA/IL-UW-961/2020\|EPI_ISL_509802\|2020-06- | ···················· | ···················· | ···················· | ···················· | ···················· | ···················· |
| F | 396 | hCoV-19/USA/IL-UW-975/2020\|EPI_ISL_509803\|2020-07- | ···················· | ···················· | ···················· | ···················· | ···················· | ···················· |
| F | 397 | hCoV-19/USA/IL-UW-979/2020\|EPI_ISL_509804\|2020-07- | ···················· | ···················· | ···················· | ···················· | ···················· | ···················· |
| F | 398 | hCoV-19/USA/IL-UW-987/2020\|EPI_ISL_509805\|2020-07- | ···················· | ···················· | ···················· | ···················· | ···················· | ···················· |
| F | 399 | hCoV-19/USA/IL-UW-996/2020\|EPI_ISL_509806\|2020-07- | ···················· | ····A··············· | ···················· | ···················· | ···················· | ···················· |
| F | 400 | hCoV-19/USA/OH-UW-921/2020\|EPI_ISL_509807\|2020-07- | ···················· | ···················· | ···················· | ···················· | ···················· | ···················· |
| F | 401 | hCoV-19/USA/WI-UW-1000/2020\|EPI_ISL_509809\|2020-07 | ···················· | ···················· | ···················· | ···················· | ···················· | ···················· |
| F | 402 | hCoV-19/USA/WI-UW-1001/2020\|EPI_ISL_509810\|2020-07 | ···················· | ·A·················· | ···················· | ···················· | ···················· | ···················· |
| F | 403 | hCoV-19/USA/WI-UW-1002/2020\|EPI_ISL_509811\|2020-07 | ···················· | ···················· | ···················· | ···················· | ···················· | ···················· |
| F | 404 | hCoV-19/USA/WI-UW-1003/2020\|EPI_ISL_509812\|2020-07 | ···················· | ·A·················· | ···················· | ···················· | ···················· | ···················· |
| F | 405 | hCoV-19/USA/WI-UW-1004/2020\|EPI_ISL_509813\|2020-07 | ···················· | ···················· | ···················· | ···················· | ···················· | ···················· |
| F | 406 | hCoV-19/USA/WI-UW-1005/2020\|EPI_ISL_509814\|2020-07 | ···················· | ···················· | ···················· | ···················· | ···················· | ···················· |
| F | 407 | hCoV-19/USA/WI-UW-806/2020\|EPI_ISL_509815\|2020-07- | ···················· | ···················· | ···················· | ···················· | ···················· | ···················· |
| F | 408 | hCoV-19/USA/WI-UW-807/2020\|EPI_ISL_509816\|2020-05- | ···················· | ···················· | ···················· | ···················· | ···················· | ···················· |
| F | 409 | hCoV-19/USA/WI-UW-808/2020\|EPI_ISL_509817\|2020-07- | ···················· | ···················· | ···················· | ···················· | ···················· | ···················· |
| F | 410 | hCoV-19/USA/WI-UW-809/2020\|EPI_ISL_509818\|2020-06- | ···················· | ···················· | ···················· | ···················· | ···················· | ···················· |
| F | 411 | hCoV-19/SaudiArabia/KAUST-JEDDAH866/2020\|EPI_ISL_5 | ···················· | ···················· | ···················· | ···················· | ···················· | ···················· |
| F | 412 | hCoV-19/SaudiArabia/KAUST-JEDDAH867/2020\|EPI_ISL_5 | ···················· | ···················· | ···················· | ···················· | ···················· | ···················· |
| F | 413 | hCoV-19/SaudiArabia/KAUST-JEDDAH868/2020\|EPI_ISL_5 | ···················· | ···················· | ···················· | ···················· | ···················· | ···················· |
| F | 414 | hCoV-19/SaudiArabia/KAUST-JEDDAH873/2020\|EPI_ISL_5 | ···················· | ···················· | ···················· | ···················· | ···················· | ···················· |
| F | 415 | hCoV-19/SaudiArabia/KAUST-JEDDAH875/2020\|EPI_ISL_5 | ···················· | ·A·················· | ···················· | ···················· | ···················· | ···A················ |
| F | 416 | hCoV-19/SaudiArabia/KAUST-JEDDAH882/2020\|EPI_ISL_5 | ···················· | ···················· | ···················· | ···················· | ···················· | ···················· |
| F | 417 | hCoV-19/SaudiArabia/KAUST-JEDDAH883/2020\|EPI_ISL_5 | ···················· | ·A·················· | ···················· | ···················· | ···················· | ···A················ |
| F | 418 | hCoV-19/SaudiArabia/KAUST-JEDDAH884/2020\|EPI_ISL_5 | ···················· | ·A·················· | ···················· | ···················· | ···················· | ···A················ |
| F | 419 | hCoV-19/SaudiArabia/KAUST-JEDDAH886/2020\|EPI_ISL_5 | ···················· | ···················· | ···················· | ···················· | ···················· | ···················· |
| F | 420 | hCoV-19/SaudiArabia/KAUST-JEDDAH887/2020\|EPI_ISL_5 | ···················· | ···················· | ···················· | ···················· | ···················· | ···················· |
| F | 421 | hCoV-19/SaudiArabia/KAUST-JEDDAH888/2020\|EPI_ISL_5 | ···················· | ···················· | ···················· | ···················· | ···················· | ···················· |
| F | 422 | hCoV-19/SaudiArabia/KAUST-JEDDAH889/2020\|EPI_ISL_5 | ···················· | ···················· | ···················· | ···················· | ···················· | ···················· |
| F | 423 | hCoV-19/SaudiArabia/KAUST-JEDDAH890/2020\|EPI_ISL_5 | ···················· | ···················· | ···················· | ···················· | ···················· | ···················· |
| F | 424 | hCoV-19/SaudiArabia/KAUST-JEDDAH891/2020\|EPI_ISL_5 | ········T··········· | ···················· | ···················· | ···················· | ···················· | ···················· |
| F | 425 | hCoV-19/Saudi Arabia/KAUST-JEDDAH900/2020\|EPI_ISL_ | ···················· | ···················· | ···················· | ···················· | ···················· | ···················· |
| F | 426 | hCoV-19/SaudiArabia/KAUST-JEDDAH903/2020\|EPI_ISL_5 | ···················· | ···················· | ···················· | ···················· | ···················· | ···················· |
| F | 427 | hCoV-19/SaudiArabia/KAUST-JEDDAH905/2020\|EPI_ISL_5 | ···················· | ···················· | ···················· | ···················· | ···················· | ···················· |
| F | 428 | hCoV-19/SaudiArabia/KAUST-JEDDAH912/2020\|EPI_ISL_5 | ···················· | ···················· | ···················· | ···················· | ···················· | ···················· |
| F | 429 | hCoV-19/SaudiArabia/KAUST-JEDDAH913/2020\|EPI_ISL_5 | ···················· | ···················· | ···················· | ···················· | ···················· | ···················· |
| F | 430 | hCoV-19/SaudiArabia/KAUST-JEDDAH916/2020\|EPI_ISL_5 | ···················· | ···················· | ···················· | ···················· | ···················· | ···················· |
| F | 431 | hCoV-19/SaudiArabia/KAUST-JEDDAH917/2020\|EPI_ISL_5 | ···················· | ···················· | ···················· | ···················· | ···················· | ···················· |
| F | 432 | hCoV-19/SaudiArabia/KAUST-JEDDAH918/2020\|EPI_ISL_5 | ···················· | ···················· | ···················· | ···················· | ···················· | ···················· |
| F | 433 | hCoV-19/SaudiArabia/KAUST-JEDDAH919/2020\|EPI_ISL_5 | ···················· | ···················· | ···················· | ···················· | ···················· | ···················· |
| F | 434 | hCoV-19/SaudiArabia/KAUST-JEDDAH920/2020\|EPI_ISL_5 | ···················· | ···················· | ···················· | ···················· | ···················· | ···················· |
| F | 435 | hCoV-19/SaudiArabia/KAUST-JEDDAH921/2020\|EPI_ISL_5 | ···················· | ···················· | ···················· | ···················· | ···················· | ···················· |
| F | 436 | hCoV-19/SaudiArabia/KAUST-JEDDAH922/2020\|EPI_ISL_5 | ···················· | ···················· | ···················· | ···················· | ···················· | ···················· |
| F | 437 | hCoV-19/SaudiArabia/KAUST-JEDDAH923/2020\|EPI_ISL_5 | ···················· | ···················· | ···················· | ···················· | ···················· | ···················· |
| F | 438 | hCoV-19/SaudiArabia/KAUST-JEDDAH926/2020\|EPI_ISL_5 | ···················· | ···················· | ···················· | ···················· | ···················· | ···················· |
| F | 439 | hCoV-19/SaudiArabia/KAUST-JEDDAH927/2020\|EPI_ISL_5 | ···················· | ···················· | ···················· | ···················· | ···················· | ···················· |
| F | 440 | hCoV-19/SaudiArabia/KAUST-JEDDAH929/2020\|EPI_ISL_5 | ···················· | ···················· | ···················· | ···················· | ···················· | ···················· |
| F | 441 | hCoV-19/SaudiArabia/KAUST-JEDDAH930/2020\|EPI_ISL_5 | ···················· | ···················· | ···················· | ···················· | ···················· | ···················· |
| F | 442 | hCoV-19/SaudiArabia/KAUST-JEDDAH931/2020\|EPI_ISL_5 | ···················· | ···················· | ···················· | ···················· | ···················· | ···················· |
| F | 443 | hCoV-19/SaudiArabia/KAUST-JEDDAH932/2020\|EPI_ISL_5 | ···················· | ···················· | ···················· | ···················· | ···················· | ···················· |
| F | 444 | hCoV-19/SaudiArabia/KAUST-JEDDAH933/2020\|EPI_ISL_5 | ···················· | ···················· | ···················· | ···················· | ···················· | ···················· |
| F | 445 | hCoV-19/SaudiArabia/KAUST-JEDDAH935/2020\|EPI_ISL_5 | ···················· | ···················· | ···················· | ···················· | ···················· | ···················· |
| F | 446 | hCoV-19/SaudiArabia/KAUST-JEDDAH936/2020\|EPI_ISL_5 | ···················· | ···················· | ···················· | ···················· | ···················· | ···················· |
| F | 447 | hCoV-19/SaudiArabia/KAUST-JEDDAH990/2020\|EPI_ISL_5 | ···················· | ···················· | ···················· | ···················· | ···················· | ···················· |
| F | 448 | hCoV-19/SaudiArabia/KAUST-JEDDAH991/2020\|EPI_ISL_5 | ···················· | ···················· | ···················· | ···················· | ···················· | ···················· |
| F | 449 | hCoV-19/SaudiArabia/KAUST-JEDDAH992/2020\|EPI_ISL_5 | ···················· | ···················· | ···················· | ···················· | ···················· | ···················· |
| F | 450 | hCoV-19/SaudiArabia/KAUST-JEDDAH993/2020\|EPI_ISL_5 | ···················· | ···················· | ···················· | ···················· | ···················· | ···················· |

| | | | | | | | |
|---|---|---|---|---|---|---|---|
| F | 451 hCoV-19/SaudiArabia/KAUST-JEDDAH996/2020\|EPI_ISL_5 | ···················· | ···················· | ···················· | ···················· | ···················· | ···················· |
| F | 452 hCoV-19/SaudiArabia/KAUST-JEDDAH997/2020\|EPI_ISL_5 | ···················· | ···················· | ···················· | ···················· | ···················· | ···················· |
| F | 453 hCoV-19/SaudiArabia/KAUST-JEDDAH998/2020\|EPI_ISL_5 | ···················· | ···················· | ···················· | ···················· | ···················· | ···················· |
| F | 454 hCoV-19/SaudiArabia/KAUST-JEDDAH999/2020\|EPI_ISL_5 | ···················· | ···················· | ···················· | ···················· | ···················· | ···················· |
| F | 455 hCoV-19/SaudiArabia/KAUST-MADINAH1036/2020\|EPI_ISL | ···················· | ···················· | ···················· | ···················· | ···················· | ···················· |
| F | 456 hCoV-19/SaudiArabia/KAUST-MADINAH1038/2020\|EPI_ISL | ···················· | ·A·················· | ···················· | ···················· | ···················· | ···A················ |
| F | 457 hCoV-19/SaudiArabia/KAUST-MADINAH1043/2020\|EPI_ISL | ···················· | ·A·················· | ···················· | ···················· | ···················· | ···A················ |
| F | 458 hCoV-19/SaudiArabia/KAUST-MADINAH1044/2020\|EPI_ISL | ···················· | ···················· | ···················· | ···················· | ···················· | ···················· |
| F | 459 hCoV-19/SaudiArabia/KAUST-MADINAH1046/2020\|EPI_ISL | ···················· | ···················· | ···················· | ···················· | ···················· | ···················· |
| F | 460 hCoV-19/SaudiArabia/KAUST-MADINAH1047/2020\|EPI_ISL | ···················· | ·A·················· | ···················· | ···················· | ···················· | ···A················ |
| F | 461 hCoV-19/SaudiArabia/KAUST-MADINAH1048/2020\|EPI_ISL | ···················· | ···················· | ···················· | ···················· | ···················· | ···················· |
| F | 462 hCoV-19/Israel/CVL-n26425/2020\|EPI_ISL_514300\|2020 | ···················· | ···················· | ···················· | ···················· | ···················· | ···················· |
| F | 463 hCoV-19/Israel/CVL-n26427/2020\|EPI_ISL_514301\|2020 | ···················· | ·A·················· | ···················· | ···················· | ···················· | ···················· |
| F | 464 hCoV-19/Israel/CVL-n26455/2020\|EPI_ISL_514302\|2020 | ···················· | ·A·················· | ···················· | ···················· | ···················· | ···················· |
| F | 466 hCoV-19/Israel/CVL-n26496/2020\|EPI_ISL_514305\|2020 | ···················· | ····A··············· | ···················· | ···················· | ···················· | ···················· |
| F | 467 hCoV-19/Israel/CVL-n26800/2020\|EPI_ISL_514306\|2020 | ···················· | ····A··············· | ···················· | ···················· | ···················· | ···················· |
| F | 468 hCoV-19/Israel/CVL-n14271/2020\|EPI_ISL_514311\|2020 | ···················· | ···················· | ···················· | ···················· | ···················· | ···················· |
| F | 469 hCoV-19/England/203290067/2020\|EPI_ISL_514350\|2020 | ···················· | ···················· | ···················· | ···················· | ···················· | ···················· |
| F | 470 hCoV-19/England/203290068/2020\|EPI_ISL_514351\|2020 | ···················· | ···················· | ···················· | ···················· | ···················· | ···················· |
| F | 471 hCoV-19/England/203290069/2020\|EPI_ISL_514352\|2020 | ···················· | ···················· | ···················· | ···················· | ···················· | ···················· |
| F | 472 hCoV-19/South Africa/LR00128/2020\|EPI_ISL_514360\|2 | ···················· | ···················· | ···················· | ···················· | ···················· | ···················· |
| F | 473 hCoV-19/South Africa/LR00136/2020\|EPI_ISL_514367\|2 | ···················· | ···················· | ···················· | ···················· | ···················· | ···················· |
| F | 474 hCoV-19/South Africa/LR00062/2020\|EPI_ISL_514375\|2 | ···················· | ···················· | ···················· | ···················· | ···················· | ···················· |
| F | 475 hCoV-19/South Africa/LR00070/2020\|EPI_ISL_514382\|2 | ···················· | ···················· | ···················· | ···················· | ···················· | ···················· |
| F | 476 hCoV-19/South Africa/KRISP-K002202/2020\|EPI_ISL_51 | ···················· | ···················· | ···················· | ···················· | ···················· | ···················· |
| F | 477 hCoV-19/South Africa/KRISP-K002203/2020\|EPI_ISL_51 | ···················· | ···················· | ···················· | ···················· | ···················· | ···················· |
| F | 478 hCoV-19/South Africa/KRISP-K002204/2020\|EPI_ISL_51 | ···················· | ···················· | ···················· | ···················· | ···················· | ···················· |
| F | 479 hCoV-19/South Africa/KRISP-K002205/2020\|EPI_ISL_51 | ···················· | ···················· | ···················· | ···················· | ···················· | ···················· |
| F | 480 hCoV-19/South Africa/KRISP-K002206/2020\|EPI_ISL_51 | ···················· | ···················· | ···················· | ···················· | ···················· | ···················· |
| F | 481 hCoV-19/South Africa/KRISP-K002209/2020\|EPI_ISL_51 | ···················· | ···················· | ···················· | ···················· | ···················· | ···················· |
| F | 482 hCoV-19/South Africa/KRISP-K002210/2020\|EPI_ISL_51 | ···················· | ···················· | ···················· | ···················· | ···················· | ···················· |
| F | 483 hCoV-19/South Africa/KRISP-K002213/2020\|EPI_ISL_51 | ···················· | ···················· | ···················· | ···················· | ···················· | ···················· |
| F | 484 hCoV-19/South Africa/KRISP-K002215/2020\|EPI_ISL_51 | ···················· | ···················· | ···················· | ···················· | ···················· | ···················· |
| F | 485 hCoV-19/South Africa/KRISP-K002216/2020\|EPI_ISL_51 | ···················· | ···················· | ···················· | ···················· | ···················· | ···················· |
| F | 486 hCoV-19/South Africa/KRISP-K002218/2020\|EPI_ISL_51 | ········T··········· | ···················· | ···················· | ···················· | ···················· | ···················· |
| F | 487 hCoV-19/South Africa/KRISP-K002219/2020\|EPI_ISL_51 | ···················· | ···················· | ···················· | ···················· | ···················· | ···················· |
| F | 488 hCoV-19/South Africa/KRISP-K002222/2020\|EPI_ISL_51 | ···················· | ···················· | ···················· | ···················· | ···················· | ···················· |
| F | 489 hCoV-19/South Africa/KRISP-K002226/2020\|EPI_ISL_51 | ···················· | ···················· | ···················· | ···················· | ···················· | ···················· |
| F | 490 hCoV-19/South Africa/KRISP-K002227/2020\|EPI_ISL_51 | ···················· | ···················· | ···················· | ···················· | ···················· | ···················· |
| F | 491 hCoV-19/South Africa/KRISP-K002231/2020\|EPI_ISL_51 | ···················· | ···················· | ···················· | ···················· | ···················· | ···················· |
| F | 492 hCoV-19/South Africa/KRISP-K002234/2020\|EPI_ISL_51 | ···················· | ···················· | ···················· | ···················· | ···················· | ···················· |
| F | 493 hCoV-19/South Africa/KRISP-K002239/2020\|EPI_ISL_51 | ···················· | ···················· | ···················· | ···················· | ···················· | ···················· |
| F | 494 hCoV-19/South Africa/KRISP-K002242/2020\|EPI_ISL_51 | ···················· | ···················· | ···················· | ···················· | ···················· | ···················· |
| F | 495 hCoV-19/South Africa/KRISP-K002243/2020\|EPI_ISL_51 | ···················· | ···················· | ···················· | ···················· | ···················· | ···················· |
| F | 497 hCoV-19/South Africa/KRISP-K002257/2020\|EPI_ISL_51 | ···················· | ···················· | ···················· | ···················· | ···················· | ···················· |
| F | 498 hCoV-19/South Africa/KRISP-K002303/2020\|EPI_ISL_51 | ···················· | ···················· | ···················· | ···················· | ···················· | ···················· |
| F | 499 hCoV-19/South Africa/KRISP-K002304/2020\|EPI_ISL_51 | ···················· | ···················· | ···················· | ···················· | ···················· | ···················· |
| F | 500 hCoV-19/South Africa/KRISP-K002306/2020\|EPI_ISL_51 | ···················· | ···················· | ···················· | ···················· | ···················· | ···················· |
| F | 501 hCoV-19/South Africa/KRISP-K002307/2020\|EPI_ISL_51 | ···················· | ···················· | ···················· | ···················· | ···················· | ···················· |
| F | 502 hCoV-19/South Africa/KRISP-K002308/2020\|EPI_ISL_51 | ···················· | ···················· | ···················· | ···················· | ···················· | ···················· |
| F | 503 hCoV-19/South Africa/KRISP-K002313/2020\|EPI_ISL_51 | ···················· | ···················· | ···················· | ···················· | ···················· | ···················· |
| F | 505 hCoV-19/South Africa/KRISP-K002315/2020\|EPI_ISL_51 | ···················· | ···················· | ···················· | ···················· | ···················· | ···················· |
| F | 506 hCoV-19/South Africa/KRISP-K002316/2020\|EPI_ISL_51 | ···················· | ···················· | ···················· | ···················· | ···················· | ···················· |
| F | 507 hCoV-19/South Africa/KRISP-K002317/2020\|EPI_ISL_51 | ···················· | ···················· | ···················· | ···················· | ···················· | ···················· |
| F | 508 hCoV-19/South Africa/KRISP-K002318/2020\|EPI_ISL_51 | ···················· | ···················· | ···················· | ···················· | ···················· | ···················· |
| F | 509 hCoV-19/South Africa/KRISP-K002320/2020\|EPI_ISL_51 | ···················· | ···················· | ···················· | ···················· | ···················· | ···················· |
| F | 510 hCoV-19/South Africa/KRISP-K002321/2020\|EPI_ISL_51 | ···················· | ···················· | ···················· | ···················· | ···················· | ···················· |
| F | 511 hCoV-19/South Africa/KRISP-K002322/2020\|EPI_ISL_51 | ···················· | ···················· | ···················· | ···················· | ···················· | ···················· |
| F | 512 hCoV-19/South Africa/KRISP-K002323/2020\|EPI_ISL_51 | ···················· | ···················· | ···················· | ···················· | ···················· | ···················· |
| F | 513 hCoV-19/Northern Ireland/NIRE-10682F/2020\|EPI_ISL_ | ···················· | ···················· | ···················· | ···················· | ···················· | ···················· |
| F | 514 hCoV-19/Northern Ireland/NIRE-10636A/2020\|EPI_ISL_ | ···················· | ···················· | ···················· | ···················· | ···················· | ···················· |
| F | 515 hCoV-19/Northern Ireland/NIRE-106F66/2020\|EPI_ISL_ | ···················· | ···················· | ···················· | ···················· | ···················· | ···················· |
| F | 516 hCoV-19/Northern Ireland/NIRE-106B62/2020\|EPI_ISL_ | ···················· | ···················· | ···················· | ···················· | ···················· | ···················· |
| F | 517 hCoV-19/Northern Ireland/NIRE-106F57/2020\|EPI_ISL_ | ···················· | ···················· | ···················· | ···················· | ···················· | ···················· |
| F | 518 hCoV-19/Northern Ireland/NIRE-106B44/2020\|EPI_ISL_ | ···················· | ···················· | ···················· | ···················· | ···················· | ···················· |
| F | 519 hCoV-19/Northern Ireland/NIRE-106D3F/2020\|EPI_ISL_ | ···················· | ···················· | ···················· | ···················· | ···················· | ···················· |
| F | 520 hCoV-19/Northern Ireland/NIRE-106C6F/2020\|EPI_ISL_ | ···················· | ···················· | ···················· | ···················· | ···················· | ···················· |
| F | 521 hCoV-19/Northern Ireland/NIRE-1065BF/2020\|EPI_ISL_ | ···················· | ···················· | ···················· | ···················· | ···················· | ···················· |
| F | 522 hCoV-19/Northern Ireland/NIRE-10635B/2020\|EPI_ISL_ | ···················· | ···················· | ···················· | ···················· | ···················· | ···················· |
| F | 523 hCoV-19/Northern Ireland/NIRE-106D6C/2020\|EPI_ISL_ | ···················· | ···················· | ···················· | ···················· | ···················· | ···················· |
| F | 524 hCoV-19/Northern Ireland/NIRE-10634C/2020\|EPI_ISL_ | ···················· | ···················· | ···················· | ···················· | ···················· | ···················· |
| F | 525 hCoV-19/Northern Ireland/NIRE-1069D1/2020\|EPI_ISL_ | ···················· | ···················· | ···················· | ···················· | ···················· | ···················· |
| F | 526 hCoV-19/Northern Ireland/NIRE-106E78/2020\|EPI_ISL_ | ···················· | ···················· | ···················· | ···················· | ···················· | ···················· |
| F | 527 hCoV-19/Northern Ireland/NIRE-1065EC/2020\|EPI_ISL_ | ···················· | ···················· | ···················· | ···················· | ···················· | ···················· |
| F | 528 hCoV-19/Northern Ireland/NIRE-106C7E/2020\|EPI_ISL_ | ···················· | ···················· | ···················· | ···················· | ···················· | ···················· |
| F | 529 hCoV-19/Northern Ireland/NIRE-106BBD/2020\|EPI_ISL_ | ···················· | ···················· | ···················· | ···················· | ···················· | ···················· |
| F | 530 hCoV-19/Northern Ireland/NIRE-106A29/2020\|EPI_ISL_ | ···················· | ···················· | ···················· | ···················· | ···················· | ···················· |
| F | 531 hCoV-19/Northern Ireland/NIRE-106AED/2020\|EPI_ISL_ | ···················· | ···················· | ···················· | ···················· | ···················· | ···················· |
| F | 532 hCoV-19/Northern Ireland/NIRE-106E1E/2020\|EPI_ISL_ | ···················· | ···················· | ···················· | ···················· | ···················· | ···················· |
| F | 533 hCoV-19/Scotland/EDB6757/2020\|EPI_ISL_507200\|2020- | ···················· | ···················A | ···················· | ···················· | ···················· | ···················· |

| | | | | | | | | |
|---|---|---|---|---|---|---|---|---|
| F | 534 hCoV-19/Scotland/EDB6760/2020\|EPI_ISL_507201\|2020- | .................... | ...................A | .................... | .................... | .................... | .................... | 1 |
| F | 535 hCoV-19/Russia/Bryansk-RII24784S/2020\|EPI_ISL_5072 | .................... | .................... | .................... | .................... | .................... | .................... | |
| F | 536 hCoV-19/Russia/Novosibirsk-RII27312S/2020\|EPI_ISL_ | .................... | .................... | .................... | .................... | .................... | .................... | |
| F | 537 hCoV-19/Russia/Novosibirsk-RII27313S/2020\|EPI_ISL_ | .................... | .................... | .................... | .................... | .................... | .................... | |
| F | 538 hCoV-19/Russia/Novosibirsk-RII27314S/2020\|EPI_ISL_ | .................... | .................... | .................... | .................... | .................... | .................... | |
| F | 539 hCoV-19/Russia/Novosibirsk-RII27315S/2020\|EPI_ISL_ | .................... | .................... | .................... | .................... | .................... | .................... | |
| F | 540 hCoV-19/Russia/Novosibirsk-RII27316S/2020\|EPI_ISL_ | .................... | .................... | .................... | .................... | .................... | .................... | |
| F | 541 hCoV-19/Russia/Novosibirsk-RII27317S/2020\|EPI_ISL_ | .................... | .................... | .................... | .................... | .................... | .................... | |
| F | 542 hCoV-19/Russia/Novosibirsk-RII27318S/2020\|EPI_ISL_ | .................... | .................... | .................... | .................... | .................... | .................... | |
| F | 543 hCoV-19/Russia/Novosibirsk-RII27320S/2020\|EPI_ISL_ | .................... | .................... | .................... | .................... | .................... | .................... | |
| F | 544 hCoV-19/Russia/Novosibirsk-RII27321S/2020\|EPI_ISL_ | .................... | .................... | .................... | .................... | .................... | .................... | |
| F | 545 hCoV-19/Russia/Novosibirsk-RII27322S/2020\|EPI_ISL_ | .................... | .................... | .................... | .................... | .................... | .................... | |
| F | 546 hCoV-19/Russia/Novy_Urengoy-RII24772S/2020\|EPI_ISL | .................... | .................... | .................... | .................... | .................... | .................... | |
| F | 547 hCoV-19/Russia/Novy_Urengoy-RII24774S/2020\|EPI_ISL | .................... | .................... | .................... | .................... | .................... | .................... | |
| F | 548 hCoV-19/Russia/Novy_Urengoy-RII24775S/2020\|EPI_ISL | .................... | .................... | .................... | .................... | .................... | .................... | |
| F | 549 hCoV-19/Russia/Novy_Urengoy-RII24776S/2020\|EPI_ISL | .................... | .................... | .................... | .................... | .................... | .................... | |
| F | 550 hCoV-19/Russia/Novy_Urengoy-RII24781S/2020\|EPI_ISL | .................... | .................... | .................... | .................... | .................... | .................... | |
| F | 551 hCoV-19/Russia/Novy_Urengoy-RII24783S/2020\|EPI_ISL | .................... | .................... | .................... | .................... | .................... | .................... | |
| F | 552 hCoV-19/Russia/Novy_Urengoy-RII24786S/2020\|EPI_ISL | .................... | .................... | .................... | .................... | .................... | .................... | |
| F | 553 hCoV-19/Russia/Novy_Urengoy-RII24787S/2020\|EPI_ISL | .................... | .................... | .................... | .................... | .................... | .................... | |
| F | 554 hCoV-19/Russia/StPetersburg-RII10066V/2020\|EPI_ISL | .................... | .................... | .................... | .................... | .................... | .................... | |
| F | 555 hCoV-19/Russia/StPetersburg-RII10128S/2020\|EPI_ISL | .................... | .................... | .................... | .................... | .................... | .................... | |
| F | 556 hCoV-19/Russia/StPetersburg-RII10183S/2020\|EPI_ISL | .................... | .................... | .................... | .................... | .................... | .................... | |
| F | 557 hCoV-19/Russia/StPetersburg-RII10219S/2020\|EPI_ISL | .................... | .................... | .................... | .................... | .................... | .................... | |
| F | 558 hCoV-19/Russia/StPetersburg-RII10223S/2020\|EPI_ISL | .................... | .................... | .................... | .................... | .................... | .................... | |
| F | 559 hCoV-19/Russia/StPetersburg-RII10261S/2020\|EPI_ISL | .................... | .................... | .................... | .................... | .................... | .................... | |
| F | 560 hCoV-19/Russia/StPetersburg-RII10289S/2020\|EPI_ISL | .................... | .................... | .................... | .................... | .................... | .................... | |
| F | 561 hCoV-19/Russia/StPetersburg-RII11126S/2020\|EPI_ISL | .................... | .................... | .................... | .................... | .................... | .................... | |
| F | 562 hCoV-19/Russia/StPetersburg-RII11142S/2020\|EPI_ISL | .................... | .................... | .................... | .................... | .................... | .................... | |
| F | 563 hCoV-19/Russia/StPetersburg-RII11144S/2020\|EPI_ISL | .................... | .................... | .................... | .................... | .................... | .................... | |
| F | 564 hCoV-19/Russia/StPetersburg-RII11149S/2020\|EPI_ISL | .................... | .................... | .................... | .................... | .................... | .................... | |
| F | 565 hCoV-19/Russia/StPetersburg-RII11149V/2020\|EPI_ISL | .................... | .................... | .................... | .................... | .................... | .................... | |
| F | 566 hCoV-19/Russia/StPetersburg-RII11151S/2020\|EPI_ISL | .................... | .................... | .................... | .................... | .................... | .................... | |
| F | 567 hCoV-19/Russia/StPetersburg-RII11153S/2020\|EPI_ISL | .................... | .................... | .................... | .................... | .................... | .................... | |
| F | 568 hCoV-19/Russia/StPetersburg-RII11155S/2020\|EPI_ISL | .................... | .A.................. | .................... | .................... | .................... | .................... | |
| F | 569 hCoV-19/Russia/StPetersburg-RII11164S/2020\|EPI_ISL | .................... | .................... | .................... | .................... | .................... | .................... | |
| F | 570 hCoV-19/USA/WI-UW-812/2020\|EPI_ISL_509820\|2020-06- | .................... | .................... | .................... | .................... | .................... | .................... | |
| F | 571 hCoV-19/USA/WI-UW-813/2020\|EPI_ISL_509821\|2020-06- | .................... | .................... | .................... | .................... | .................... | .................... | |
| F | 572 hCoV-19/USA/WI-UW-815/2020\|EPI_ISL_509822\|2020-06- | .................... | .................... | .................... | .................... | .................... | .................... | |
| F | 573 hCoV-19/USA/WI-UW-816/2020\|EPI_ISL_509823\|2020-07- | .................... | .................... | .................... | .................... | .................... | .................... | |
| F | 574 hCoV-19/USA/WI-UW-817/2020\|EPI_ISL_509824\|2020-05- | .................... | .................... | .................... | .................... | .................... | .................... | |
| F | 575 hCoV-19/USA/WI-UW-818/2020\|EPI_ISL_509825\|2020-05- | .................... | .................... | .................... | .................... | .................... | .................... | |
| F | 576 hCoV-19/USA/WI-UW-819/2020\|EPI_ISL_509826\|2020-05- | .................... | .................... | .................... | .................... | .................... | .................... | |
| F | 577 hCoV-19/USA/WI-UW-820/2020\|EPI_ISL_509827\|2020-05- | .................... | .................... | .................... | .................... | .................... | .................... | |
| F | 578 hCoV-19/USA/WI-UW-821/2020\|EPI_ISL_509828\|2020-05- | .................... | .................... | .................... | .................... | .................... | .................... | |
| F | 579 hCoV-19/USA/WI-UW-822/2020\|EPI_ISL_509829\|2020-05- | .................... | .................... | .................... | .................... | .................... | .................... | |
| F | 580 hCoV-19/USA/WI-UW-833/2020\|EPI_ISL_509840\|2020-05- | .................... | .................... | .................... | .................... | .................... | .................... | |
| F | 581 hCoV-19/USA/WI-UW-834/2020\|EPI_ISL_509841\|2020-05- | .................... | .................... | .................... | .................... | .................... | .................... | |
| F | 582 hCoV-19/USA/WI-UW-835/2020\|EPI_ISL_509842\|2020-05- | .................... | .................... | .................... | .................... | .................... | .................... | |
| F | 583 hCoV-19/USA/WI-UW-836/2020\|EPI_ISL_509843\|2020-05- | .................... | .................... | .................... | .................... | .................... | .................... | |
| F | 584 hCoV-19/USA/WI-UW-837/2020\|EPI_ISL_509844\|2020-05- | .................... | .................... | .................... | .................... | .................... | .................... | |
| F | 585 hCoV-19/USA/WI-UW-838/2020\|EPI_ISL_509845\|2020-05- | .................... | .................... | .................... | .................... | .................... | .................... | |
| F | 586 hCoV-19/USA/WI-UW-840/2020\|EPI_ISL_509847\|2020-05- | .................... | .................... | .................... | .................... | .................... | .................... | |
| F | 588 hCoV-19/USA/WI-UW-842/2020\|EPI_ISL_509849\|2020-05- | .................... | .................... | .................... | .................... | .................... | .................... | |
| F | 589 hCoV-19/USA/WI-UW-843/2020\|EPI_ISL_509850\|2020-05- | .................... | .................... | .................... | .................... | .................... | .................... | |
| F | 590 hCoV-19/USA/WI-UW-844/2020\|EPI_ISL_509851\|2020-05- | .................... | .................... | .................... | .................... | .................... | .................... | |
| F | 592 hCoV-19/USA/WI-UW-847/2020\|EPI_ISL_509853\|2020-07- | .................... | .................... | .................... | .................... | .................... | .................... | |
| F | 593 hCoV-19/USA/WI-UW-848/2020\|EPI_ISL_509854\|2020-06- | .................... | .................... | .................... | .................... | .................... | .................... | |
| F | 594 hCoV-19/USA/WI-UW-849/2020\|EPI_ISL_509855\|2020-07- | .................... | .................... | .................... | .................... | .................... | .................... | |
| F | 595 hCoV-19/USA/WI-UW-850/2020\|EPI_ISL_509856\|2020-07- | .................... | .................... | .................... | .................... | .................... | .................... | |
| F | 596 hCoV-19/USA/WI-UW-851/2020\|EPI_ISL_509857\|2020-07- | .................... | .................... | .................... | .................... | .................... | .................... | |
| F | 599 hCoV-19/USA/WI-UW-855/2020\|EPI_ISL_509861\|2020-07- | .................... | .................... | .................... | .................... | .................... | .................... | |
| F | 604 hCoV-19/USA/WI-UW-864/2020\|EPI_ISL_509867\|2020-07- | .................... | .................... | .................... | .................... | .................... | .................... | |
| F | 605 hCoV-19/USA/WI-UW-865/2020\|EPI_ISL_509868\|2020-07- | .................... | .................... | .................... | .................... | .................... | .................... | |
| F | 606 hCoV-19/USA/WI-UW-866/2020\|EPI_ISL_509869\|2020-07- | .................... | .................... | .................... | .................... | .................... | .................... | |
| F | 607 hCoV-19/USA/WI-UW-867/2020\|EPI_ISL_509870\|2020-07- | .................... | .................... | .................... | .................... | .................... | .................... | |
| F | 608 hCoV-19/USA/WI-UW-868/2020\|EPI_ISL_509871\|2020-07- | .................... | .................... | .................... | .................... | .................... | .................... | |
| F | 609 hCoV-19/USA/WI-UW-870/2020\|EPI_ISL_509872\|2020-07- | .................... | .................... | .................... | .................... | .................... | .................... | |
| F | 610 hCoV-19/USA/WI-UW-872/2020\|EPI_ISL_509874\|2020-07- | .................... | .................... | .................... | .................... | .................... | .................... | |
| F | 611 hCoV-19/USA/WI-UW-873/2020\|EPI_ISL_509875\|2020-07- | .................... | .................... | .................... | .................... | .................... | .................... | |
| F | 612 hCoV-19/USA/WI-UW-874/2020\|EPI_ISL_509876\|2020-07- | .................... | .................... | .................... | .................... | .................... | .................... | |
| F | 613 hCoV-19/USA/WI-UW-875/2020\|EPI_ISL_509877\|2020-07- | .................... | .................... | .................... | .................... | .................... | .................... | |
| F | 614 hCoV-19/USA/WI-UW-876/2020\|EPI_ISL_509878\|2020-07- | .................... | .................... | .................... | .................... | .................... | .................... | |
| F | 615 hCoV-19/USA/WI-UW-877/2020\|EPI_ISL_509879\|2020-07- | .................... | .................... | .................... | .................... | .................... | .................... | |
| F | 616 hCoV-19/USA/WI-UW-878/2020\|EPI_ISL_509880\|2020-07- | .................... | .................... | .................... | .................... | .................... | .................... | |
| F | 617 hCoV-19/USA/WI-UW-879/2020\|EPI_ISL_509881\|2020-07- | .................... | .................... | .................... | .................... | .................... | .................... | |
| F | 618 hCoV-19/USA/WI-UW-880/2020\|EPI_ISL_509882\|2020-07- | .................... | .................... | .................... | .................... | .................... | .................... | |
| F | 619 hCoV-19/USA/WI-UW-881/2020\|EPI_ISL_509883\|2020-07- | .................... | .A.................. | .................... | .................... | .................... | .................... | |
| F | 620 hCoV-19/USA/WI-UW-882/2020\|EPI_ISL_509884\|2020-07- | .................... | .................... | .................... | .................... | .................... | .................... | |
| F | 621 hCoV-19/USA/WI-UW-883/2020\|EPI_ISL_509885\|2020-07- | .................... | .................... | .................... | .................... | .................... | .................... | |

| | | | | | | | | |
|---|---|---|---|---|---|---|---|---|
| F | 622 hCoV-19/USA/WI-UW-884/2020\|EPI_ISL_509886\|2020-07- | ···················· | ···················· | ····················· | ···················· | ···················· | ···················· | |
| F | 623 hCoV-19/USA/WI-UW-886/2020\|EPI_ISL_509888\|2020-07- | ···················· | ···················· | ····················· | ···················· | ···················· | ···················· | |
| F | 624 hCoV-19/USA/WI-UW-887/2020\|EPI_ISL_509889\|2020-07- | ···················· | ···················· | ····················· | ···················· | ···················· | ···················· | |
| F | 625 hCoV-19/USA/WI-UW-888/2020\|EPI_ISL_509890\|2020-07- | ···················· | ···················· | ····················· | ···················· | ···················· | ···················· | |
| F | 626 hCoV-19/USA/WI-UW-889/2020\|EPI_ISL_509891\|2020-07- | ···················· | ···················· | ····················· | ···················· | ···················· | ···················· | |
| F | 627 hCoV-19/USA/WI-UW-890/2020\|EPI_ISL_509892\|2020-07- | ···················· | ···················· | ····················· | ···················· | ···················· | ···················· | |
| F | 628 hCoV-19/USA/WI-UW-891/2020\|EPI_ISL_509893\|2020-07- | ···················· | ···················· | ····················· | ···················· | ···················· | ···················· | |
| F | 629 hCoV-19/USA/WI-UW-892/2020\|EPI_ISL_509894\|2020-07- | ···················· | ···················· | ····················· | ···················· | ···················· | ···················· | |
| F | 630 hCoV-19/USA/WI-UW-893/2020\|EPI_ISL_509895\|2020-07- | ···················· | ···················· | ····················· | ···················· | ···················· | ···················· | |
| F | 631 hCoV-19/USA/WI-UW-894/2020\|EPI_ISL_509896\|2020-07- | ···················· | ···················· | ····················· | ···················· | ···················· | ···················· | |
| F | 632 hCoV-19/USA/WI-UW-895/2020\|EPI_ISL_509897\|2020-07- | ···················· | ···················· | ····················· | ···················· | ···················· | ···················· | |
| F | 633 hCoV-19/USA/WI-UW-896/2020\|EPI_ISL_509898\|2020-07- | ···················· | ···················· | ····················· | ···················· | ···················· | ···················· | |
| F | 634 hCoV-19/USA/WI-UW-897/2020\|EPI_ISL_509899\|2020-07- | ···················· | ···················· | ····················· | ···················· | ···················· | ···················· | |
| F | 635 hCoV-19/Sweden/20-08902/2020\|EPI_ISL_510810\|2020-0 | ···················· | ···················· | ····················· | ···················· | ···················· | ···················· | |
| F | 636 hCoV-19/Sweden/20-08904/2020\|EPI_ISL_510811\|2020-0 | ···················· | ···················· | ····················· | ···················· | ···················· | ···················· | |
| F | 637 hCoV-19/Sweden/20-08905/2020\|EPI_ISL_510812\|2020-0 | ···················· | ···················· | ····················· | ···················· | ···················· | ···················· | |
| F | 638 hCoV-19/Sweden/20-08906/2020\|EPI_ISL_510813\|2020-0 | ···················· | ···················· | ····················· | ···················· | ···················· | ···················· | |
| F | 639 hCoV-19/Sweden/20-08908/2020\|EPI_ISL_510814\|2020-0 | ···················· | ···················· | ····················· | ···················· | ···················· | ···················· | |
| F | 640 hCoV-19/Sweden/20-08909/2020\|EPI_ISL_510815\|2020-0 | ···················· | ···················· | ····················· | ···················· | ···················· | ···················· | |
| F | 641 hCoV-19/Sweden/20-08910/2020\|EPI_ISL_510816\|2020-0 | ···················· | ···················· | ····················· | ···················· | ···················· | ···················· | |
| F | 642 hCoV-19/Sweden/20-50310/2020\|EPI_ISL_510817\|2020-0 | ···················· | ···················· | ····················· | ···················· | ···················· | ···················· | |
| F | 643 hCoV-19/Sweden/20-50564/2020\|EPI_ISL_510818\|2020-0 | ···················· | ···················· | ····················· | ···················· | ···················· | ···················· | |
| F | 644 hCoV-19/Sweden/20-50934/2020\|EPI_ISL_510819\|2020-0 | ···················· | ···················· | ····················· | ···················· | ···················· | ···················· | |
| F | 645 hCoV-19/Sweden/20-51272/2020\|EPI_ISL_510820\|2020-0 | ···················· | ···················· | ····················· | ···················· | ···················· | ···················· | |
| F | 646 hCoV-19/Sweden/20-51275/2020\|EPI_ISL_510821\|2020-0 | ···················· | ···················· | ····················· | ···················· | ···················· | ···················· | |
| F | 647 hCoV-19/Sweden/20-51277/2020\|EPI_ISL_510822\|2020-0 | ···················· | ···················· | ····················· | ···················· | ···················· | ···················· | |
| F | 648 hCoV-19/Sweden/20-51278/2020\|EPI_ISL_510823\|2020-0 | ···················· | ···················· | ····················· | ···················· | ···················· | ···················· | |
| F | 649 hCoV-19/Sweden/20-51279/2020\|EPI_ISL_510824\|2020-0 | ···················· | ···················· | ····················· | ···················· | ···················· | ···················· | |
| F | 650 hCoV-19/Sweden/20-51416/2020\|EPI_ISL_510825\|2020-0 | ···················· | ···················· | ····················· | ···················· | ···················· | ···················· | |
| F | 651 hCoV-19/Sweden/20-51417/2020\|EPI_ISL_510826\|2020-0 | ···················· | ···················· | ····················· | ···················· | ···················· | ···················· | |
| F | 652 hCoV-19/Sweden/20-51531/2020\|EPI_ISL_510827\|2020-0 | ···················· | ···················· | ····················· | ···················· | ···················· | ···················· | |
| F | 653 hCoV-19/Sweden/20-51534/2020\|EPI_ISL_510828\|2020-0 | ···················· | ···················· | ····················· | ···················· | ···················· | ···················· | |
| F | 654 hCoV-19/Sweden/20-51536/2020\|EPI_ISL_510829\|2020-0 | ···················· | ···················· | ····················· | ···················· | ···················· | ···················· | |
| F | 655 hCoV-19/Sweden/20-51538/2020\|EPI_ISL_510830\|2020-0 | ···················· | ···················· | ····················· | ···················· | ···················· | ···················· | |
| F | 656 hCoV-19/Sweden/20-51570/2020\|EPI_ISL_510831\|2020-0 | ···················· | ···················· | ····················· | ···················· | ···················· | ···················· | |
| F | 657 hCoV-19/Sweden/20-51576/2020\|EPI_ISL_510832\|2020-0 | ···················· | ···················· | ····················· | ···················· | ···················· | ···················· | |
| F | 658 hCoV-19/Sweden/20-51579/2020\|EPI_ISL_510833\|2020-0 | ···················· | ···················· | ····················· | ···················· | ···················· | ···················· | |
| F | 659 hCoV-19/Sweden/20-51697/2020\|EPI_ISL_510834\|2020-0 | ···················· | ···················· | ····················· | ···················· | ···················· | ···················· | |
| F | 660 hCoV-19/Sweden/20-51698/2020\|EPI_ISL_510835\|2020-0 | ···················· | ···················· | ····················· | ···················· | ···················· | ···················· | |
| F | 661 hCoV-19/Sweden/20-51710/2020\|EPI_ISL_510836\|2020-0 | ···················· | ···················· | ····················· | ···················· | ···················· | ···················· | |
| F | 662 hCoV-19/Sweden/20-51711/2020\|EPI_ISL_510837\|2020-0 | ···················· | ···················· | ····················· | ···················· | ···················· | ···················· | |
| F | 663 hCoV-19/Sweden/20-51733/2020\|EPI_ISL_510838\|2020-0 | ···················· | ···················· | ····················· | ···················· | ···················· | ···················· | |
| F | 664 hCoV-19/Sweden/20-51736/2020\|EPI_ISL_510839\|2020-0 | ···················· | ···················· | ····················· | ···················· | ···················· | ···················· | |
| F | 665 hCoV-19/Sweden/20-51737/2020\|EPI_ISL_510840\|2020-0 | ···················· | ···················· | ····················· | ···················· | ···················· | ···················· | |
| F | 666 hCoV-19/Sweden/20-51740/2020\|EPI_ISL_510841\|2020-0 | ···················· | ···················· | ····················· | ···················· | ···················· | ···················· | |
| F | 667 hCoV-19/Sweden/20-51745/2020\|EPI_ISL_510842\|2020-0 | ···················· | ···················· | ····················· | ···················· | ···················· | ···················· | |
| F | 668 hCoV-19/Sweden/20-51746/2020\|EPI_ISL_510843\|2020-0 | ···················· | ···················· | ··················T·· | ···················· | ···················· | ···················· | 1 |
| F | 669 hCoV-19/Sweden/20-51748/2020\|EPI_ISL_510844\|2020-0 | ···················· | ···················· | ····················· | ···················· | ···················· | ···················· | |
| F | 670 hCoV-19/Sweden/20-51765/2020\|EPI_ISL_510845\|2020-0 | ···················· | ···················· | ····················· | ···················· | ···················· | ···················· | |
| F | 671 hCoV-19/Sweden/20-51767/2020\|EPI_ISL_510846\|2020-0 | ···················· | ···················· | ····················· | ···················· | ···················· | ···················· | |
| F | 672 hCoV-19/Sweden/20-51769/2020\|EPI_ISL_510847\|2020-0 | ···················· | ···················· | ····················· | ···················· | ···················· | ···················· | |
| F | 673 hCoV-19/Sweden/20-51771/2020\|EPI_ISL_510848\|2020-0 | ···················· | ···················· | ····················· | ···················· | ···················· | ···················· | |
| F | 674 hCoV-19/Sweden/20-51772/2020\|EPI_ISL_510849\|2020-0 | ···················· | ···················· | ····················· | ···················· | ···················· | ···················· | |
| F | 675 hCoV-19/Sweden/20-51773/2020\|EPI_ISL_510850\|2020-0 | ···················· | ···················· | ····················· | ···················· | ···················· | ···················· | |
| F | 676 hCoV-19/Sweden/20-51774/2020\|EPI_ISL_510851\|2020-0 | ···················· | ···················· | ····················· | ···················· | ···················· | ···················· | |
| F | 677 hCoV-19/Sweden/20-51844/2020\|EPI_ISL_510852\|2020-0 | ···················· | ···················· | ····················· | ···················· | ···················· | ···················· | |
| F | 678 hCoV-19/Sweden/20-51847/2020\|EPI_ISL_510853\|2020-0 | ···················· | ···················· | ····················· | ···················· | ···················· | ···················· | |
| F | 679 hCoV-19/Sweden/20-51848/2020\|EPI_ISL_510854\|2020-0 | ···················· | ···················· | ····················· | ···················· | ···················· | ···················· | |
| F | 680 hCoV-19/Sweden/20-51849/2020\|EPI_ISL_510855\|2020-0 | ···················· | ···················· | ····················· | ···················· | ···················· | ···················· | |
| F | 681 hCoV-19/Sweden/20-51852/2020\|EPI_ISL_510856\|2020-0 | ···················· | ···················· | ····················· | ···················· | ···················· | ···················· | |
| F | 682 hCoV-19/Sweden/20-51853/2020\|EPI_ISL_510857\|2020-0 | ···················· | ···················· | ····················· | ···················· | ···················· | ···················· | |
| F | 683 hCoV-19/Sweden/20-51854/2020\|EPI_ISL_510858\|2020-0 | ···················· | ···················· | ····················· | ···················· | ···················· | ···················· | |
| F | 684 hCoV-19/Sweden/20-51894/2020\|EPI_ISL_510859\|2020-0 | ···················· | ···················· | ····················· | ···················· | ···················· | ···················· | |
| F | 685 hCoV-19/Sweden/20-51946/2020\|EPI_ISL_510860\|2020-0 | ···················· | ···················· | ····················· | ···················· | ···················· | ···················· | |
| F | 686 hCoV-19/Sweden/20-51947/2020\|EPI_ISL_510861\|2020-0 | ···················· | ·A·················· | ····················· | ···················· | ···················· | ···················· | |
| F | 687 hCoV-19/Sweden/20-51948/2020\|EPI_ISL_510862\|2020-0 | ···················· | ···················· | ····················· | ···················· | ···················· | ···················· | |
| F | 688 hCoV-19/Sweden/20-51949/2020\|EPI_ISL_510863\|2020-0 | ···················· | ···················· | ····················· | ···················· | ···················· | ···················· | |
| F | 689 hCoV-19/Sweden/20-51997/2020\|EPI_ISL_510864\|2020-0 | ···················· | ···················· | ····················· | ···················· | ···················· | ···················· | |
| F | 690 hCoV-19/Sweden/20-52000/2020\|EPI_ISL_510865\|2020-0 | ···················· | ···················· | ····················· | ···················· | ···················· | ···················· | |
| F | 691 hCoV-19/Sweden/20-52025/2020\|EPI_ISL_510866\|2020-0 | ···················· | ···················· | ····················· | ···················· | ···················· | ···················· | |
| F | 692 hCoV-19/Sweden/20-52036/2020\|EPI_ISL_510867\|2020-0 | ···················· | ···················· | ····················· | ···················· | ···················· | ···················· | |
| F | 693 hCoV-19/Sweden/20-52048/2020\|EPI_ISL_510868\|2020-0 | ···················· | ···················· | ····················· | ···················· | ···················· | ···················· | |
| F | 694 hCoV-19/Sweden/20-52049/2020\|EPI_ISL_510869\|2020-0 | ···················· | ···················· | ····················· | ···················· | ···················· | ···················· | |
| F | 695 hCoV-19/Sweden/20-52052/2020\|EPI_ISL_510870\|2020-0 | ···················· | ···················· | ··················T·· | ···················· | ···················· | ···················· | 1 |
| F | 696 hCoV-19/Sweden/20-52071/2020\|EPI_ISL_510871\|2020-0 | ···················· | ···················· | ····················· | ···················· | ···················· | ···················· | |
| F | 697 hCoV-19/Sweden/20-52073/2020\|EPI_ISL_510872\|2020-0 | ···················· | ···················· | ····················· | ···················· | ···················· | ···················· | |
| F | 698 hCoV-19/Sweden/20-52074/2020\|EPI_ISL_510873\|2020-0 | ···················· | ···················· | ····················· | ···················· | ···················· | ···················· | |
| F | 699 hCoV-19/Sweden/20-52077/2020\|EPI_ISL_510874\|2020-0 | ···················· | ···················· | ····················· | ···················· | ···················· | ···················· | |
| F | 700 hCoV-19/Sweden/20-52078/2020\|EPI_ISL_510875\|2020-0 | ···················· | ···················· | ····················· | ···················· | ···················· | ···················· | |
| F | 701 hCoV-19/Sweden/20-52079/2020\|EPI_ISL_510876\|2020-0 | ···················· | ···················· | ····················· | ···················· | ···················· | ···················· | |

| | | | | | | | |
|---|---|---|---|---|---|---|---|
| F | 702 hCoV-19/Sweden/20-52080/2020\|EPI_ISL_510877\|2020-0 | ···················· | ···················· | ····················· | ···················· | ···················· | ···················· |
| F | 703 hCoV-19/USA/NM-NMDOH-2020242033/2020\|EPI_ISL_51088 | ···················· | ·A·················· | ····················· | ···················· | ···················· | ···················· |
| F | 704 hCoV-19/USA/NM-NMDOH-2020242034/2020\|EPI_ISL_51088 | ···················· | ·A·················· | ····················· | ···················· | ···················· | ···················· |
| F | 705 hCoV-19/USA/NM-NMDOH-2020242035/2020\|EPI_ISL_51088 | ···················· | ·A·················· | ····················· | ···················· | ···················· | ···················· |
| F | 706 hCoV-19/USA/NM-NMDOH-2020242037/2020\|EPI_ISL_51088 | ···················· | ·A·················· | ····················· | ···················· | ···················· | ···················· |
| F | 707 hCoV-19/SaudiArabia/KAUST-JEDDAH757/2020\|EPI_ISL_5 | ···················· | ···················· | ····················· | ···················· | ···················· | ···················· |
| F | 708 hCoV-19/SaudiArabia/KAUST-JEDDAH759/2020\|EPI_ISL_5 | ···················· | ···················· | ····················· | ···················· | ···················· | ···················· |
| F | 709 hCoV-19/SaudiArabia/KAUST-JEDDAH826/2020\|EPI_ISL_5 | ···················· | ···················· | ····················· | ···················· | ···················· | ···················· |
| F | 710 hCoV-19/SaudiArabia/KAUST-JEDDAH828/2020\|EPI_ISL_5 | ···················· | ···················· | ····················· | ···················· | ···················· | ···················· |
| F | 711 hCoV-19/SaudiArabia/KAUST-JEDDAH830/2020\|EPI_ISL_5 | ···················· | ···················· | ····················· | ···················· | ···················· | ···················· |
| F | 712 hCoV-19/SaudiArabia/KAUST-JEDDAH831/2020\|EPI_ISL_5 | ···················· | ···················· | ····················· | ···················· | ···················· | ···················· |
| F | 713 hCoV-19/SaudiArabia/KAUST-JEDDAH833/2020\|EPI_ISL_5 | ···················· | ···················· | ····················· | ···················· | ···················· | ···················· |
| F | 714 hCoV-19/SaudiArabia/KAUST-JEDDAH835/2020\|EPI_ISL_5 | ···················· | ···················· | ····················· | ···················· | ···················· | ···················· |
| F | 715 hCoV-19/SaudiArabia/KAUST-JEDDAH836/2020\|EPI_ISL_5 | ···················· | ···················· | ····················· | ···················· | ···················· | ···················· |
| F | 716 hCoV-19/SaudiArabia/KAUST-JEDDAH837/2020\|EPI_ISL_5 | ···················· | ···················· | ····················· | ···················· | ···················· | ···················· |
| F | 717 hCoV-19/SaudiArabia/KAUST-JEDDAH840/2020\|EPI_ISL_5 | ···················· | ···················· | ····················· | ···················· | ···················· | ···················· |
| F | 718 hCoV-19/SaudiArabia/KAUST-JEDDAH841/2020\|EPI_ISL_5 | ···················· | ···················· | ····················· | ···················· | ···················· | ···················· |
| F | 719 hCoV-19/SaudiArabia/KAUST-JEDDAH842/2020\|EPI_ISL_5 | ···················· | ···················· | ····················· | ···················· | ···················· | ···················· |
| F | 720 hCoV-19/SaudiArabia/KAUST-JEDDAH843/2020\|EPI_ISL_5 | ···················· | ···················· | ····················· | ···················· | ···················· | ···················· |
| F | 721 hCoV-19/SaudiArabia/KAUST-JEDDAH844/2020\|EPI_ISL_5 | ···················· | ···················· | ····················· | ···················· | ···················· | ···················· |
| F | 722 hCoV-19/SaudiArabia/KAUST-JEDDAH845/2020\|EPI_ISL_5 | ···················· | ···················· | ····················· | ···················· | ···················· | ···················· |
| F | 723 hCoV-19/SaudiArabia/KAUST-JEDDAH846/2020\|EPI_ISL_5 | ···················· | ···················· | ····················· | ···················· | ···················· | ···················· |
| F | 724 hCoV-19/SaudiArabia/KAUST-JEDDAH847/2020\|EPI_ISL_5 | ···················· | ···················· | ····················· | ···················· | ···················· | ···················· |
| F | 725 hCoV-19/SaudiArabia/KAUST-JEDDAH869/2020\|EPI_ISL_5 | ···················· | ···················· | ····················· | ···················· | ···················· | ···················· |
| F | 726 hCoV-19/SaudiArabia/KAUST-JEDDAH870/2020\|EPI_ISL_5 | ···················· | ···················· | ····················· | ···················· | ···················· | ···················· |
| F | 727 hCoV-19/SaudiArabia/KAUST-JEDDAH871/2020\|EPI_ISL_5 | ···················· | ···················· | ····················· | ···················· | ···················· | ···················· |
| F | 728 hCoV-19/SaudiArabia/KAUST-JEDDAH872/2020\|EPI_ISL_5 | ···················· | ···················· | ····················· | ···················· | ···················· | ···················· |
| F | 729 hCoV-19/SaudiArabia/KAUST-JEDDAH877/2020\|EPI_ISL_5 | ···················· | ···················· | ····················· | ···················· | ···················· | ···················· |
| F | 730 hCoV-19/SaudiArabia/KAUST-MADINAH1073/2020\|EPI_ISL | ···················· | ·A·················· | ····················· | ···················· | ···················· | ··A················· |
| F | 731 hCoV-19/SaudiArabia/KAUST-MADINAH1074/2020\|EPI_ISL | ···················· | ·A·················· | ····················· | ···················· | ···················· | ··A················· |
| F | 732 hCoV-19/SaudiArabia/KAUST-MADINAH1076/2020\|EPI_ISL | ···················· | ·A·················· | ····················· | ···················· | ···················· | ··A················· |
| F | 733 hCoV-19/SaudiArabia/KAUST-MADINAH1079/2020\|EPI_ISL | ···················· | ···················· | ····················· | ···················· | ···················· | ···················· |
| F | 734 hCoV-19/SaudiArabia/KAUST-MADINAH1081/2020\|EPI_ISL | ···················· | ···················· | ····················· | ···················· | ···················· | ···················· |
| F | 735 hCoV-19/SaudiArabia/KAUST-MADINAH1085/2020\|EPI_ISL | ···················· | ·A·················· | ····················· | ···················· | ···················· | ··A················· |
| F | 736 hCoV-19/SaudiArabia/KAUST-MADINAH1086/2020\|EPI_ISL | ···················· | ·A·················· | ····················· | ···················· | ···················· | ··A················· |
| F | 737 hCoV-19/Peru/CUS-INS-065/2020\|EPI_ISL_514338\|2020- | ···················· | ···················· | ····················· | ···················· | ···················· | ···················· |
| F | 738 hCoV-19/Peru/LOR-INS-079/2020\|EPI_ISL_514339\|2020- | ···················· | ···················· | ····················· | ···················· | ···················· | ···················· |
| F | 739 hCoV-19/Peru/LOR-INS-080/2020\|EPI_ISL_514340\|2020- | ···················· | ···················· | ····················· | ···················· | ···················· | ···················· |
| F | 740 hCoV-19/Peru/LOR-INS-081/2020\|EPI_ISL_514341\|2020- | ···················· | ···················· | ····················· | ···················· | ···················· | ···················· |
| F | 741 hCoV-19/Peru/LOR-INS-082/2020\|EPI_ISL_514342\|2020- | ···················· | ···················· | ····················· | ···················· | ···················· | ···················· |
| F | 742 hCoV-19/England/203290059/2020\|EPI_ISL_514343\|2020 | ···················· | ···················· | ····················· | ···················· | ···················· | ···················· |
| F | 743 hCoV-19/England/203290060/2020\|EPI_ISL_514344\|2020 | ···················· | ···················· | ····················· | ···················· | ···················· | ···················· |
| F | 744 hCoV-19/England/203290061/2020\|EPI_ISL_514345\|2020 | ···················· | ···················· | ····················· | ···················· | ···················· | ···················· |
| F | 745 hCoV-19/England/203290063/2020\|EPI_ISL_514346\|2020 | ···················· | ···················· | ····················· | ···················· | ···················· | ···················· |
| F | 746 hCoV-19/England/203290064/2020\|EPI_ISL_514347\|2020 | ···················· | ···················· | ····················· | ···················· | ···················· | ···················· |
| F | 747 hCoV-19/England/203290065/2020\|EPI_ISL_514348\|2020 | ···················· | ···················· | ····················· | ···················· | ···················· | ···················· |
| F | 748 hCoV-19/England/203290066/2020\|EPI_ISL_514349\|2020 | ···················· | ···················· | ····················· | ···················· | ···················· | ···················· |
| F | 749 hCoV-19/North Macedonia/2175/2020\|EPI_ISL_514354\|2 | ···················· | ···················· | ····················· | ···················· | ···················· | ···················· |
| F | 750 hCoV-19/South Africa/LR00122/2020\|EPI_ISL_514355\|2 | ···················· | ···················· | ····················· | ···················· | ···················· | ···················· |
| F | 751 hCoV-19/South Africa/KRISP-K002280/2020\|EPI_ISL_51 | ···················· | ···················· | ····················· | ···················· | ···················· | ···················· |
| F | 752 hCoV-19/South Africa/KRISP-K002281/2020\|EPI_ISL_51 | ···················· | ···················· | ····················· | ···················· | ···················· | ···················· |
| F | 753 hCoV-19/South Africa/KRISP-K002282/2020\|EPI_ISL_51 | ···················· | ···················· | ····················· | ···················· | ···················· | ···················· |
| F | 754 hCoV-19/South Africa/KRISP-K002287/2020\|EPI_ISL_51 | ···················· | ···················· | ····················· | ···················· | ···················· | ···················· |
| F | 755 hCoV-19/South Africa/KRISP-K002288/2020\|EPI_ISL_51 | ···················· | ···················· | ····················· | ···················· | ···················· | ···················· |
| F | 756 hCoV-19/South Africa/KRISP-K002290/2020\|EPI_ISL_51 | ···················· | ···················· | ····················· | ···················· | ···················· | ···················· |
| F | 757 hCoV-19/South Africa/KRISP-K002291/2020\|EPI_ISL_51 | ···················· | ···················· | ····················· | ···················· | ···················· | ···················· |
| F | 758 hCoV-19/South Africa/KRISP-K002293/2020\|EPI_ISL_51 | ···················· | ···················· | ····················· | ···················· | ···················· | ···················· |
| F | 759 hCoV-19/South Africa/KRISP-K002295/2020\|EPI_ISL_51 | ···················· | ···················· | ····················· | ···················· | ···················· | ···················· |
| F | 760 hCoV-19/South Africa/KRISP-K002297/2020\|EPI_ISL_51 | ···················· | ···················· | ····················· | ···················· | ···················· | ···················· |
| F | 761 hCoV-19/South Africa/KRISP-K002298/2020\|EPI_ISL_51 | ···················· | ···················· | ····················· | ···················· | ···················· | ···················· |
| F | 762 hCoV-19/South Africa/KRISP-K002299/2020\|EPI_ISL_51 | ···················· | ···················· | ····················· | ···················· | ···················· | ···················· |
| F | 763 hCoV-19/South Africa/KRISP-K002301/2020\|EPI_ISL_51 | ···················· | ···················· | ····················· | ···················· | ···················· | ···················· |
| F | 764 hCoV-19/South Africa/KRISP-K002302/2020\|EPI_ISL_51 | ···················· | ···················· | ····················· | ···················· | ···················· | ···················· |
| F | 767 hCoV-19/South Africa/KRISP-K002311/2020\|EPI_ISL_51 | ···················· | ···················· | ····················· | ···················· | ···················· | ···················· |
| F | 768 hCoV-19/South Africa/KRISP-K002312/2020\|EPI_ISL_51 | ···················· | ···················· | ····················· | ···················· | ···················· | ···················· |
| F | 769 hCoV-19/USA/WA-S1610/2020\|EPI_ISL_496036\|2020-05-2 | ···················· | ···················· | ····················· | ···················· | ···················· | ···················· |
| F | 770 hCoV-19/USA/WA-S1611/2020\|EPI_ISL_496037\|2020-05-2 | ···················· | ···················· | ····················· | ···················· | ···················· | ···················· |
| F | 771 hCoV-19/USA/WA-S1612/2020\|EPI_ISL_496038\|2020-05-2 | ···················· | ···················· | ····················· | ···················· | ···················· | ···················· |
| F | 772 hCoV-19/USA/WA-S1613/2020\|EPI_ISL_496039\|2020-05-2 | ···················· | ···················· | ····················· | ···················· | ···················· | ···················· |
| F | 773 hCoV-19/USA/WA-S1614/2020\|EPI_ISL_496040\|2020-05-2 | ···················· | ···················· | ····················· | ···················· | ···················· | ···················· |
| F | 774 hCoV-19/USA/WA-S1616/2020\|EPI_ISL_496042\|2020-05-2 | ···················· | ···················· | ····················· | ···················· | ···················· | ···················· |
| F | 775 hCoV-19/USA/WA-S1619/2020\|EPI_ISL_496045\|2020-05-2 | ···················· | ···················· | ····················· | ···················· | ···················· | ···················· |
| F | 776 hCoV-19/USA/WA-S1620/2020\|EPI_ISL_496046\|2020-05-2 | ···················· | ···················· | ····················· | ···················· | ···················· | ···················· |
| F | 777 hCoV-19/USA/WA-S1621/2020\|EPI_ISL_496047\|2020-05-2 | ···················· | ···················· | ····················· | ···················· | ···················· | ···················· |
| F | 778 hCoV-19/USA/WA-S1622/2020\|EPI_ISL_496048\|2020-05-2 | ···················· | ···················· | ····················· | ···················· | ···················· | ···················· |
| F | 779 hCoV-19/USA/WA-S1623/2020\|EPI_ISL_496049\|2020-05-2 | ···················· | ···················· | ····················· | ···················· | ···················· | ···················· |
| F | 780 hCoV-19/USA/WA-S1624/2020\|EPI_ISL_496050\|2020-05-2 | ···················· | ···················· | ····················· | ···················· | ···················· | ···················· |
| F | 781 hCoV-19/USA/WA-S1625/2020\|EPI_ISL_496051\|2020-05-2 | ···················· | ···················· | ····················· | ···················· | ···················· | ···················· |
| F | 782 hCoV-19/USA/WA-S1626/2020\|EPI_ISL_496052\|2020-05-2 | ···················· | ···················· | ····················· | ···················· | ···················· | ···················· |
| F | 783 hCoV-19/USA/WA-S1627/2020\|EPI_ISL_496053\|2020-05-2 | ···················· | ···················· | ····················· | ···················· | ···················· | ···················· |

F 784 hCoV-19/USA/WA-S1628/2020|EPI_ISL_496054|2020-05-2 ···················· ···················· ····················· ···················· ···················· ····················
F 785 hCoV-19/USA/WA-S1629/2020|EPI_ISL_496055|2020-05-2 ···················· ···················· ····················· ···················· ···················· ····················
F 786 hCoV-19/USA/WA-S1630/2020|EPI_ISL_496056|2020-05-2 ···················· ···················· ····················· ···················· ···················· ····················
F 787 hCoV-19/USA/WA-S1631/2020|EPI_ISL_496057|2020-05-2 ···················· ···················· ····················· ···················· ···················· ····················
F 788 hCoV-19/USA/WA-S1654/2020|EPI_ISL_496080|2020-05-2 ···················· ···················· ····················· ···················· ···················· ····················
F 789 hCoV-19/USA/WA-S1655/2020|EPI_ISL_496081|2020-05-2 ···················· ···················· ····················· ···················· ···················· ····················
F 790 hCoV-19/USA/WA-S1656/2020|EPI_ISL_496082|2020-05-2 ···················· ···················· ····················· ···················· ···················· ····················
F 791 hCoV-19/USA/WA-S1657/2020|EPI_ISL_496083|2020-05-2 ···················· ···················· ····················· ···················· ···················· ····················
F 792 hCoV-19/USA/WA-S1658/2020|EPI_ISL_496084|2020-05-2 ···················· ···················· ····················· ···················· ···················· ····················
F 793 hCoV-19/USA/WA-S1659/2020|EPI_ISL_496085|2020-05-2 ···················· ···················· ····················· ···················· ···················· ····················
F 794 hCoV-19/USA/WA-S1660/2020|EPI_ISL_496086|2020-05-2 ···················· ···················· ····················· ···················· ···················· ····················
F 795 hCoV-19/USA/WA-S1661/2020|EPI_ISL_496087|2020-05-2 ···················· ···················· ····················· ···················· ···················· ····················
F 796 hCoV-19/USA/WA-S1662/2020|EPI_ISL_496088|2020-05-2 ···················· ···················· ····················· ···················· ···················· ····················
F 797 hCoV-19/USA/WA-S1663/2020|EPI_ISL_496089|2020-05-2 ···················· ···················· ····················· ···················· ···················· ····················
F 798 hCoV-19/USA/WA-S1664/2020|EPI_ISL_496090|2020-05-2 ···················· ···················· ····················· ···················· ···················· ····················
F 799 hCoV-19/USA/WA-S1665/2020|EPI_ISL_496091|2020-05-2 ···················· ···················· ····················· ···················· ···················· ····················
F 800 hCoV-19/USA/WA-S1666/2020|EPI_ISL_496092|2020-05-2 ···················· ···················· ····················· ···················· ···················· ····················
F 801 hCoV-19/USA/WA-S1668/2020|EPI_ISL_496094|2020-05-2 ···················· ···················· ····················· ···················· ···················· ····················
F 802 hCoV-19/USA/WA-S1670/2020|EPI_ISL_496096|2020-05-2 ···················· ···················· ····················· ···················· ···················· ····················
F 803 hCoV-19/USA/WA-S1671/2020|EPI_ISL_496097|2020-05-3 ···················· ···················· ····················· ···················· ···················· ····················
F 804 hCoV-19/USA/WA-S1672/2020|EPI_ISL_496098|2020-05-2 ···················· ···················· ····················· ···················· ···················· ····················
F 805 hCoV-19/USA/WA-S1673/2020|EPI_ISL_496099|2020-05-2 ···················· ···················· ····················· ···················· ···················· ····················
F 806 hCoV-19/USA/WA-S1939/2020|EPI_ISL_497367|2020-06-0 ···················· ···················· ····················· ···················· ···················· ····················
F 807 hCoV-19/USA/WA-S1940/2020|EPI_ISL_497368|2020-06-0 ···················· ···················· ····················· ···················· ···················· ····················
F 808 hCoV-19/USA/WA-S1941/2020|EPI_ISL_497369|2020-06-0 ···················· ···················· ····················· ···················· ···················· ····················
F 809 hCoV-19/USA/WA-S1942/2020|EPI_ISL_497370|2020-06-0 ···················· ···················· ····················· ···················· ···················· ····················
F 810 hCoV-19/USA/WA-S1943/2020|EPI_ISL_497371|2020-06-0 ···················· ···················· ····················· ···················· ···················· ····················
F 811 hCoV-19/USA/WA-S1944/2020|EPI_ISL_497372|2020-06-0 ···················· ···················· ····················· ···················· ···················· ····················
F 812 hCoV-19/USA/WA-S1945/2020|EPI_ISL_497373|2020-06-0 ···················· ···················· ····················· ···················· ···················· ····················
F 813 hCoV-19/USA/WA-S1948/2020|EPI_ISL_497376|2020-06-0 ···················· ···················· ····················· ···················· ···················· ····················
F 814 hCoV-19/USA/WA-S1949/2020|EPI_ISL_497377|2020-06-0 ···················· ···················· ····················· ···················· ···················· ····················
F 815 hCoV-19/USA/WA-S1950/2020|EPI_ISL_497378|2020-06-0 ···················· ···················· ····················· ···················· ···················· ····················
F 816 hCoV-19/USA/WA-S1951/2020|EPI_ISL_497379|2020-06-0 ···················· ···················· ····················· ···················· ···················· ····················
F 817 hCoV-19/USA/WA-S1952/2020|EPI_ISL_497380|2020-06-0 ···················· ···················· ····················· ···················· ···················· ····················
F 818 hCoV-19/USA/WA-S1953/2020|EPI_ISL_497381|2020-06-0 ···················· ···················· ····················· ···················· ···················· ····················
F 819 hCoV-19/USA/WA-S1954/2020|EPI_ISL_497382|2020-06-0 ···················· ···················· ····················· ···················· ···················· ····················
F 820 hCoV-19/USA/WA-S1955/2020|EPI_ISL_497383|2020-06-0 ···················· ···················· ····················· ···················· ···················· ····················
F 821 hCoV-19/USA/WA-S1956/2020|EPI_ISL_497384|2020-06-0 ···················· ···················· ····················· ···················· ···················· ····················
F 822 hCoV-19/USA/WA-S1957/2020|EPI_ISL_497385|2020-06-0 ···················· ···················· ····················· ···················· ···················· ····················
F 823 hCoV-19/USA/WA-S1958/2020|EPI_ISL_497386|2020-06-0 ···················· ···················· ····················· ···················· ···················· ····················
F 824 hCoV-19/USA/WA-S1959/2020|EPI_ISL_497387|2020-06-0 ···················· ···················· ····················· ···················· ···················· ····················
F 825 hCoV-19/USA/WA-S1960/2020|EPI_ISL_497388|2020-06-0 ···················· ···A··············· ····················· ···················· ···················· ····················
F 826 hCoV-19/USA/CA-QDX-222/2020|EPI_ISL_498698|2020-04 ···················· ···················· ····················· ···················· ···················· ····················
F 827 hCoV-19/USA/CA-QDX-223/2020|EPI_ISL_498699|2020-04 ···················· ···················· ····················· ···················· ···················· ····················
F 828 hCoV-19/Saudi Arabia/KAIMRC15/2020|EPI_ISL_507020| ···················· ···················· ····················· ···················· ···················· ····················
F 829 hCoV-19/Saudi Arabia/KAIMRC16/2020|EPI_ISL_507021| ···················· ···················· ····················· ···················· ···················· ····················
F 830 hCoV-19/Saudi Arabia/KAIMRC17/2020|EPI_ISL_507022| ···················· ···················· ····················· ···················· ···················· ····················
F 831 hCoV-19/Saudi Arabia/KAIMRC18/2020|EPI_ISL_507023| ···················· ···················· ····················· ···················· ···················· ····················
F 832 hCoV-19/Saudi Arabia/KAIMRC59/2020|EPI_ISL_507024| ···················· ···················· ····················· ···················· ···················· ····················
F 833 hCoV-19/Saudi Arabia/KAIMRC60/2020|EPI_ISL_507025| ···················· ···················· ····················· ···················· ···················· ····················
F 834 hCoV-19/Saudi Arabia/KAIMRC62/2020|EPI_ISL_507026| ···················· ···················· ····················· ···················· ···················· ····················
F 835 hCoV-19/Saudi Arabia/KAIMRC63/2020|EPI_ISL_507027| ···················· ···················· ····················· ···················· ···················· ····················
F 836 hCoV-19/Saudi Arabia/KAIMRC64/2020|EPI_ISL_507028| ···················· ···················· ····················· ···················· ···················· ····················
F 837 hCoV-19/Saudi Arabia/KAIMRC67/2020|EPI_ISL_507029| ···················· ···················· ····················· ···················· ···················· ····················
F 838 hCoV-19/Saudi Arabia/KAIMRC69/2020|EPI_ISL_507030| ···················· ···················· ····················· ···················· ···················· ····················
F 839 hCoV-19/Saudi Arabia/KAIMRC70/2020|EPI_ISL_507031| ···················· ···················· ····················· ···················· ···················· ····················
F 840 hCoV-19/Saudi Arabia/KFSHRC27/2020|EPI_ISL_507032| ···················· ···················· ····················· ···················· ···················· ····················
F 841 hCoV-19/Saudi Arabia/KFSHRC28/2020|EPI_ISL_507033| ···················· ···················· ····················· ···················· ···················· ····················
F 842 hCoV-19/Saudi Arabia/KFSHRC31/2020|EPI_ISL_507034| ···················· ···················· ····················· ···················· ···················· ····················
F 843 hCoV-19/Saudi Arabia/KFSHRC42/2020|EPI_ISL_507035| ···················· ···················· ····················· ···················· ···················· ····················
F 844 hCoV-19/Saudi Arabia/KFSHRC43/2020|EPI_ISL_507036| ···················· ···················· ····················· ···················· ···················· ····················
F 845 hCoV-19/Saudi Arabia/KFSHRC44/2020|EPI_ISL_507037| ···················· ···················· ····················· ···················· ···················· ····················
F 846 hCoV-19/Saudi Arabia/KFSHRC50/2020|EPI_ISL_507038| ···················· ···················· ····················· ···················· ···················· ····················
F 847 hCoV-19/South Korea/CBNU-nCoV01/2020|EPI_ISL_50703 ···················· ···················· ····················· ···················· ···················· ····················
F 848 hCoV-19/England/LOND-124CD1B/2020|EPI_ISL_507040|2 ···················· ···················· ····················· ···················· ···················· ····················
F 849 hCoV-19/England/LOND-124CD48/2020|EPI_ISL_507041|2 ···················· ···················· ····················· ···················· ···················· ····················
F 850 hCoV-19/England/LOND-124CD57/2020|EPI_ISL_507042|2 ···················· ···················· ····················· ···················· ···················· ····················
F 851 hCoV-19/England/LOND-124CDB1/2020|EPI_ISL_507043|2 ···················· ···················· ····················· ···················· ···················· ····················
F 852 hCoV-19/England/LOND-124CF8E/2020|EPI_ISL_507047|2 ···················· ···················· ····················· ···················· ···················· ····················
F 853 hCoV-19/England/LOND-124D020/2020|EPI_ISL_507048|2 ···················· ···················· ····················· ···················· ···················· ····················
F 854 hCoV-19/England/LOND-124D05D/2020|EPI_ISL_507049|2 ···················· ···················· ····················· ···················· ···················· ····················
F 855 hCoV-19/England/LOND-124D08A/2020|EPI_ISL_507050|2 ···················· ···················· ····················· ···················· ···················· ····················
F 856 hCoV-19/England/LOND-124D0A8/2020|EPI_ISL_507052|2 ···················· ···················· ····················· ···················· ···················· ····················
F 857 hCoV-19/England/LOND-124D0C6/2020|EPI_ISL_507053|2 ···················· ···················· ····················· ···················· ···················· ····················
F 858 hCoV-19/England/LOND-124D0D5/2020|EPI_ISL_507054|2 ···················· ···················· ····················· ···················· ···················· ····················
F 859 hCoV-19/England/LOND-124D0E4/2020|EPI_ISL_507055|2 ···················· ···················· ····················· ···················· ···················· ····················
F 860 hCoV-19/England/LOND-124D0F3/2020|EPI_ISL_507056|2 ···················· ···················· ····················· ···················· ···················· ····················
F 861 hCoV-19/England/LOND-124D14B/2020|EPI_ISL_507057|2 ···················· ···················· ····················· ···················· ···················· ····················
F 862 hCoV-19/England/LOND-124D15A/2020|EPI_ISL_507058|2 ···················· ···················· ····················· ···················· ···················· ····················
F 863 hCoV-19/England/LOND-124D169/2020|EPI_ISL_507059|2 ···················· ···················· ····················· ···················· ···················· ····················

| | | | | | | | |
|---|---|---|---|---|---|---|---|
| F | 864 hCoV-19/England/LOND-124D178/2020\|EPI_ISL_507060\|2 | ···················· | ···················· | ···················· | ···················· | ···················· | ···················· |
| F | 865 hCoV-19/England/LOND-124D1A5/2020\|EPI_ISL_507062\|2 | ···················· | ···················· | ···················· | ···················· | ···················· | ···················· |
| F | 866 hCoV-19/England/LOND-124D1B4/2020\|EPI_ISL_507063\|2 | ···················· | ···················· | ···················· | ···················· | ···················· | ···················· |
| F | 867 hCoV-19/England/LOND-124D1C3/2020\|EPI_ISL_507064\|2 | ···················· | ···················· | ···················· | ···················· | ···················· | ···················· |
| F | 868 hCoV-19/England/LOND-124D1D2/2020\|EPI_ISL_507065\|2 | ···················· | ···················· | ···················· | ···················· | ···················· | ···················· |
| F | 869 hCoV-19/England/LOND-124D1E1/2020\|EPI_ISL_507066\|2 | ···················· | ···················· | ···················· | ···················· | ···················· | ···················· |
| F | 870 hCoV-19/England/LOND-124D1F0/2020\|EPI_ISL_507067\|2 | ···················· | ···················· | ···················· | ···················· | ···················· | ···················· |
| F | 871 hCoV-19/England/LOND-124D20C/2020\|EPI_ISL_507068\|2 | ···················· | ···················· | ···················· | ···················· | ···················· | ···················· |
| F | 872 hCoV-19/England/LOND-124D21B/2020\|EPI_ISL_507069\|2 | ···················· | ···················· | ···················· | ···················· | ···················· | ···················· |
| F | 873 hCoV-19/England/LOND-124D239/2020\|EPI_ISL_507070\|2 | ···················· | ···················· | ···················· | ···················· | ···················· | ···················· |
| F | 874 hCoV-19/England/LOND-124D248/2020\|EPI_ISL_507071\|2 | ···················· | ···················· | ···················· | ···················· | ···················· | ···················· |
| F | 875 hCoV-19/England/LOND-124D257/2020\|EPI_ISL_507072\|2 | ···················· | ···················· | ···················· | ···················· | ···················· | ···················· |
| F | 876 hCoV-19/England/LOND-124D266/2020\|EPI_ISL_507073\|2 | ···················· | ···················· | ···················· | ···················· | ···················· | ···················· |
| F | 877 hCoV-19/England/LOND-124D275/2020\|EPI_ISL_507074\|2 | ···················· | ···················· | ···················· | ···················· | ···················· | ···················· |
| F | 878 hCoV-19/England/LOND-124D284/2020\|EPI_ISL_507075\|2 | ···················· | ···················· | ···················· | ···················· | ···················· | ···················· |
| F | 879 hCoV-19/England/LOND-124D293/2020\|EPI_ISL_507076\|2 | ···················· | ···················· | ···················· | ···················· | ···················· | ···················· |
| F | 880 hCoV-19/England/LOND-124D2A2/2020\|EPI_ISL_507077\|2 | ···················· | ···················· | ···················· | ···················· | ···················· | ···················· |
| F | 881 hCoV-19/England/LOND-124D2B1/2020\|EPI_ISL_507078\|2 | ···················· | ···················· | ···················· | ···················· | ···················· | ···················· |
| F | 882 hCoV-19/England/LOND-124D2C0/2020\|EPI_ISL_507079\|2 | ···················· | ···················· | ···················· | ···················· | ···················· | ···················· |
| F | 883 hCoV-19/India/WB-IK20/2020\|EPI_ISL_508350\|2020-04- | ···················· | ···················· | ···················· | ···················· | ···················· | ···················· |
| F | 884 hCoV-19/India/WB-IK21/2020\|EPI_ISL_508351\|2020-04- | ···················· | ···················· | ···················· | ···················· | ···················· | ···················· |
| F | 885 hCoV-19/India/WB-IK22/2020\|EPI_ISL_508352\|2020-04- | ···················· | ···················· | ···················· | ···················· | ···················· | ···················· |
| F | 886 hCoV-19/India/WB-IK23/2020\|EPI_ISL_508353\|2020-04- | ···················· | ···················· | ···················· | ···················· | ···················· | ···················· |
| F | 887 hCoV-19/India/WB-IK24/2020\|EPI_ISL_508354\|2020-04- | ···················· | ···················· | ···················· | ···················· | ···················· | ···················· |
| F | 888 hCoV-19/India/WB-IK25/2020\|EPI_ISL_508355\|2020-04- | ···················· | ···················· | ···················· | ···················· | ···················· | ···················· |
| F | 889 hCoV-19/India/WB-IK26/2020\|EPI_ISL_508356\|2020-04- | ···················· | ···················· | ···················· | ···················· | ···················· | ···················· |
| F | 890 hCoV-19/India/WB-IK27/2020\|EPI_ISL_508357\|2020-04- | ···················· | ···················· | ···················· | ···················· | ···················· | ···················· |
| F | 891 hCoV-19/India/WB-IK28/2020\|EPI_ISL_508358\|2020-04- | ···················· | ···················· | ···················· | ···················· | ···················· | ···················· |
| F | 892 hCoV-19/India/WB-IK29/2020\|EPI_ISL_508359\|2020-04- | ···················· | ···················· | ···················· | ···················· | ···················· | ···················· |
| F | 893 hCoV-19/India/WB-IK30/2020\|EPI_ISL_508360\|2020-04- | ···················· | ···················· | ···················· | ···················· | ··A················· | ···················· |
| F | 894 hCoV-19/India/WB-IK31/2020\|EPI_ISL_508361\|2020-04- | ···················· | ···················· | ···················· | ···················· | ··A················· | ···················· |
| F | 895 hCoV-19/India/WB-IK32/2020\|EPI_ISL_508362\|2020-05- | ···················· | ···················· | ···················· | ···················· | ···················· | ···················· |
| F | 896 hCoV-19/India/WB-IK33/2020\|EPI_ISL_508363\|2020-05- | ···················· | ···················· | ···················· | ···················· | ···················· | ···················· |
| F | 897 hCoV-19/India/WB-IK35/2020\|EPI_ISL_508364\|2020-05- | ···················· | ···················· | ···················· | ···················· | ···················· | ···················· |
| F | 898 hCoV-19/India/WB-IK36/2020\|EPI_ISL_508365\|2020-05- | ···················· | ···················· | ···················· | ···················· | ···················· | ···················· |
| F | 899 hCoV-19/India/WB-IK38/2020\|EPI_ISL_508366\|2020-05- | ···················· | ···················· | ···················· | ···················· | ···················· | ···················· |
| F | 900 hCoV-19/India/WB-IK39/2020\|EPI_ISL_508367\|2020-05- | ···················· | ···················· | ···················· | ···················· | ···················· | ···················· |
| F | 901 hCoV-19/India/WB-IK4/2020\|EPI_ISL_508368\|2020-03-3 | ···················· | ···················· | ···················· | ···················· | ···················· | ···················· |
| F | 902 hCoV-19/India/WB-IK41/2020\|EPI_ISL_508369\|2020-05- | ···················· | ···················· | ···················· | ···················· | ···················· | ···················· |
| F | 903 hCoV-19/India/WB-IK42/2020\|EPI_ISL_508370\|2020-05- | ···················· | ···················· | ···················· | ···················· | ···················· | ···················· |
| F | 904 hCoV-19/India/WB-IK44/2020\|EPI_ISL_508371\|2020-05- | ···················· | ···················· | ···················· | ···················· | ···················· | ···················· |
| F | 905 hCoV-19/India/WB-IK45/2020\|EPI_ISL_508372\|2020-05- | ···················· | ···················· | ···················· | ···················· | ···················· | ···················· |
| F | 906 hCoV-19/India/WB-IK46/2020\|EPI_ISL_508373\|2020-05- | ···················· | ···················· | ···················· | ···················· | ···················· | ···················· |
| F | 907 hCoV-19/India/WB-IK47/2020\|EPI_ISL_508374\|2020-05- | ···················· | ···················· | ···················· | ···················· | ···················· | ···················· |
| F | 908 hCoV-19/India/WB-IK48/2020\|EPI_ISL_508375\|2020-05- | ···················· | ···················· | ···················· | ···················· | ···················· | ···················· |
| F | 909 hCoV-19/India/WB-IK5/2020\|EPI_ISL_508376\|2020-03-3 | ···················· | ···················· | ···················· | ···················· | ···················· | ···················· |
| F | 910 hCoV-19/India/WB-IK50/2020\|EPI_ISL_508377\|2020-05- | ···················· | ···················· | ···················· | ···················· | ···················· | ···················· |
| F | 911 hCoV-19/India/WB-IK51/2020\|EPI_ISL_508378\|2020-05- | ···················· | ···················· | ···················· | ···················· | ···················· | ···················· |
| F | 912 hCoV-19/India/WB-IK53/2020\|EPI_ISL_508379\|2020-05- | ···················· | ···················· | ···················· | ···················· | ···················· | ···················· |
| F | 913 hCoV-19/India/WB-IK54/2020\|EPI_ISL_508380\|2020-05- | ···················· | ···················· | ···················· | ···················· | ···················· | ···················· |
| F | 914 hCoV-19/India/WB-IK55/2020\|EPI_ISL_508381\|2020-05- | ···················· | ···················· | ···················· | ···················· | ···················· | ···················· |
| F | 915 hCoV-19/India/WB-IK56/2020\|EPI_ISL_508382\|2020-05- | ···················· | ···················· | ···················· | ···················· | ···················· | ···················· |
| F | 916 hCoV-19/India/WB-IK57/2020\|EPI_ISL_508383\|2020-05- | ···················· | ···················· | ···················· | ···················· | ···················· | ···················· |
| F | 917 hCoV-19/India/WB-IK58/2020\|EPI_ISL_508384\|2020-05- | ···················· | ···················· | ···················· | ···················· | ···················· | ···················· |
| F | 918 hCoV-19/India/WB-IK6/2020\|EPI_ISL_508385\|2020-04-0 | ···················· | ···················· | ···················· | ···················· | ···················· | ···················· |
| F | 919 hCoV-19/India/WB-IK60/2020\|EPI_ISL_508386\|2020-05- | ···················· | ···················· | ···················· | ···················· | ···················· | ···················· |
| F | 920 hCoV-19/India/WB-IK63/2020\|EPI_ISL_508387\|2020-05- | ···················· | ···················· | ···················· | ···················· | ···················· | ···················· |
| F | 921 hCoV-19/India/WB-IK64/2020\|EPI_ISL_508388\|2020-05- | ···················· | ···················· | ···················· | ···················· | ···················· | ···················· |
| F | 922 hCoV-19/India/WB-IK65/2020\|EPI_ISL_508389\|2020-05- | ···················· | ···················· | ···················· | ···················· | ···················· | ···················· |
| F | 923 hCoV-19/India/WB-IK67/2020\|EPI_ISL_508391\|2020-05- | ···················· | ···················· | ···················· | ···················· | ··A················· | ···················· |
| F | 924 hCoV-19/India/WB-IK68/2020\|EPI_ISL_508392\|2020-05- | ···················· | ···················· | ···················· | ···················· | ···················· | ···················· |
| F | 925 hCoV-19/India/WB-IK69/2020\|EPI_ISL_508393\|2020-05- | ···················· | ···················· | ···················· | ···················· | ···················· | ···················· |
| F | 926 hCoV-19/India/WB-IK7/2020\|EPI_ISL_508394\|2020-04-0 | ···················· | ···················· | ···················· | ···················· | ···················· | ···················· |
| F | 927 hCoV-19/India/WB-IK71/2020\|EPI_ISL_508395\|2020-05- | ···················· | ···················· | ···················· | ···················· | ···················· | ···················· |
| F | 928 hCoV-19/India/WB-IK72/2020\|EPI_ISL_508396\|2020-05- | ···················· | ···················· | ···················· | ···················· | ···················· | ···················· |
| F | 929 hCoV-19/India/WB-IK73/2020\|EPI_ISL_508397\|2020-05- | ···················· | ···················· | ···················· | ···················· | ··A················· | ···················· |
| F | 930 hCoV-19/India/WB-IK75/2020\|EPI_ISL_508398\|2020-05- | ···················· | ···················· | ···················· | ···················· | ···················· | ···················· |
| F | 931 hCoV-19/India/WB-IK77/2020\|EPI_ISL_508399\|2020-05- | ···················· | ···················· | ···················· | ···················· | ···················· | ···················· |
| F | 932 hCoV-19/India/HR-TF26/2020\|EPI_ISL_508500\|2020-06- | ···················· | ·A·················· | ···················· | ···················· | ···················· | ··A················· |
| F | 933 hCoV-19/India/HR-TF27/2020\|EPI_ISL_508501\|2020-06- | ···················· | ·A·················· | ···················· | ···················· | ···················· | ··A················· |
| F | 934 hCoV-19/India/HR-TF3/2020\|EPI_ISL_508502\|2020-06-1 | ···················· | ···················· | ···················· | ···················· | ···················· | ···················· |
| F | 935 hCoV-19/India/HR-TF30/2020\|EPI_ISL_508503\|2020-06- | ···················· | ·A·················· | ···················· | ···················· | ···················· | ··A················· |
| F | 936 hCoV-19/India/HR-TF38/2020\|EPI_ISL_508504\|2020-05- | ···················· | ···················· | ···················· | ···················· | ···················· | ···················· |
| F | 937 hCoV-19/India/HR-TF4/2020\|EPI_ISL_508505\|2020-06-1 | ···················· | ·A·················· | ···················· | ···················· | ···················· | ··A················· |
| F | 938 hCoV-19/India/HR-TF41/2020\|EPI_ISL_508506\|2020-05- | ···················· | ·A·················· | ···················· | ···················· | ···················· | ··A················· |
| F | 939 hCoV-19/India/HR-TF6/2020\|EPI_ISL_508507\|2020-06-1 | ···················· | ·A·················· | ···················· | ···················· | ···················· | ··A················· |
| F | 940 hCoV-19/India/HR-TF7/2020\|EPI_ISL_508508\|2020-06-1 | ···················· | ·A·················· | ···················· | ···················· | ···················· | ··A················· |
| F | 941 hCoV-19/India/HR-TF9/2020\|EPI_ISL_508509\|2020-06-1 | ···················· | ·A·················· | ···················· | ···················· | ···················· | ··A················· |
| F | 942 hCoV-19/USA/AR-CDC-4172/2020\|EPI_ISL_509680\|2020-0 | ···················· | ···················· | ···················· | ···················· | ···················· | ···················· |
| F | 943 hCoV-19/USA/AR-CDC-4176/2020\|EPI_ISL_509681\|2020-0 | ···················· | ···················· | ···················· | ···················· | ···················· | ···················· |

| | | | | | | | |
|---|---|---|---|---|---|---|---|
| F | 944 hCoV-19/USA/AR-CDC-4177/2020\|EPI_ISL_509682\|2020-0 | ···················· | ···················· | ···················· | ···················· | ···················· | ···················· |
| F | 945 hCoV-19/USA/AR-CDC-4181/2020\|EPI_ISL_509683\|2020-0 | ···················· | ···················· | ···················· | ···················· | ···················· | ···················· |
| F | 946 hCoV-19/USA/AR-CDC-4194/2020\|EPI_ISL_509684\|2020-0 | ···················· | ···················· | ···················· | ···················· | ···················· | ···················· |
| F | 947 hCoV-19/USA/AR-CDC-4204/2020\|EPI_ISL_509685\|2020-0 | ···················· | ···················· | ···················· | ···················· | ···················· | ···················· |
| F | 948 hCoV-19/USA/MN-MDH-1385/2020\|EPI_ISL_509686\|2020-0 | ···················· | ···················· | ···················· | ···················· | ···················· | ···················· |
| F | 949 hCoV-19/USA/AL-CDC-0731/2020\|EPI_ISL_509688\|2020-0 | ···················· | ···················· | ···················· | ···················· | ···················· | ···················· |
| F | 950 hCoV-19/USA/UT-CDC-9758/2020\|EPI_ISL_509689\|2020-0 | ···················· | ···················· | ···················· | ···················· | ···················· | ···················· |
| F | 951 hCoV-19/USA/UT-CDC-9749/2020\|EPI_ISL_509690\|2020-0 | ···················· | ···················· | ···················· | ···················· | ···················· | ···················· |
| F | 952 hCoV-19/USA/UT-CDC-9750/2020\|EPI_ISL_509691\|2020-0 | ···················· | ···················· | ···················· | ···················· | ···················· | ···················· |
| F | 953 hCoV-19/USA/UT-CDC-9755/2020\|EPI_ISL_509692\|2020-0 | ···················· | ···················· | ···················· | ···················· | ···················· | ···················· |
| F | 954 hCoV-19/USA/UT-CDC-9756/2020\|EPI_ISL_509693\|2020-0 | ···················· | ···················· | ···················· | ···················· | ···················· | ···················· |
| F | 955 hCoV-19/USA/UT-CDC-9757/2020\|EPI_ISL_509694\|2020-0 | ···················· | ···················· | ···················· | ···················· | ···················· | ···················· |
| F | 956 hCoV-19/Guatemala/CDC-1227/2020\|EPI_ISL_509695\|202 | ···················· | ···················· | ···················· | ···················· | ···················· | ···················· |
| F | 957 hCoV-19/Guatemala/CDC-1228/2020\|EPI_ISL_509696\|202 | ···················· | ···················· | ···················· | ···················· | ···················· | ···················· |
| F | 958 hCoV-19/Guatemala/CDC-1230/2020\|EPI_ISL_509697\|202 | ···················· | ···················· | ···················· | ···················· | ···················· | ···················· |
| F | 959 hCoV-19/Guatemala/CDC-1231/2020\|EPI_ISL_509698\|202 | ···················· | ···················· | ···················· | ···················· | ···················· | ···················· |
| F | 960 hCoV-19/Guatemala/CDC-1232/2020\|EPI_ISL_509699\|202 | ···················· | ···················· | ···················· | ···················· | ···················· | ···················· |
| F | 961 hCoV-19/USA/WI-UW-810/2020\|EPI_ISL_509819\|2020-06- | ···················· | ···················· | ···················· | ···················· | ···················· | ···················· |
| F | 962 hCoV-19/USA/WI-UW-823/2020\|EPI_ISL_509830\|2020-06- | ···················· | ···················· | ···················· | ···················· | ···················· | ···················· |
| F | 963 hCoV-19/USA/WI-UW-824/2020\|EPI_ISL_509831\|2020-06- | ···················· | ···················· | ···················· | ···················· | ···················· | ···················· |
| F | 964 hCoV-19/USA/WI-UW-825/2020\|EPI_ISL_509832\|2020-06- | ···················· | ···················· | ···················· | ···················· | ···················· | ···················· |
| F | 965 hCoV-19/USA/WI-UW-826/2020\|EPI_ISL_509833\|2020-06- | ···················· | ···················· | ···················· | ···················· | ···················· | ···················· |
| F | 966 hCoV-19/USA/WI-UW-827/2020\|EPI_ISL_509834\|2020-05- | ···················· | ···················· | ···················· | ···················· | ···················· | ···················· |
| F | 967 hCoV-19/USA/WI-UW-828/2020\|EPI_ISL_509835\|2020-05- | ···················· | ···················· | ···················· | ···················· | ···················· | ···················· |
| F | 968 hCoV-19/USA/WI-UW-829/2020\|EPI_ISL_509836\|2020-05- | ···················· | ···················· | ···················· | ···················· | ···················· | ···················· |
| F | 969 hCoV-19/USA/WI-UW-830/2020\|EPI_ISL_509837\|2020-05- | ···················· | ···················· | ···················· | ···················· | ···················· | ···················· |
| F | 970 hCoV-19/USA/WI-UW-831/2020\|EPI_ISL_509838\|2020-05- | ···················· | ···················· | ···················· | ···················· | ···················· | ···················· |
| F | 971 hCoV-19/USA/WI-UW-832/2020\|EPI_ISL_509839\|2020-05- | ···················· | ···················· | ···················· | ···················· | ···················· | ···················· |
| F | 972 hCoV-19/South Korea/KCDC2330/2020\|EPI_ISL_510666\|2 | ···················· | ···················· | ···················· | ···················· | ···················· | ···················· |
| F | 973 hCoV-19/South Korea/KCDC2331/2020\|EPI_ISL_510667\|2 | ···················· | ···················· | ···················· | ···················· | ···················· | ···················· |
| F | 974 hCoV-19/South Korea/KCDC2332/2020\|EPI_ISL_510668\|2 | ···················· | ···················· | ···················· | ···················· | ···················· | ···················· |
| F | 975 hCoV-19/South Korea/KCDC2333/2020\|EPI_ISL_510669\|2 | ···················· | ···················· | ···················· | ···················· | ···················· | ···················· |
| F | 976 hCoV-19/South Korea/KCDC2334/2020\|EPI_ISL_510670\|2 | ···················· | ···················· | ···················· | ···················· | ···················· | ···················· |
| F | 977 hCoV-19/South Korea/KCDC2335/2020\|EPI_ISL_510671\|2 | ···················· | ···················· | ···················· | ···················· | ···················· | ···················· |
| F | 978 hCoV-19/South Korea/KCDC2337/2020\|EPI_ISL_510673\|2 | ···················· | ···················· | ···················· | ···················· | ···················· | ···················· |
| F | 979 hCoV-19/South Korea/KCDC2338/2020\|EPI_ISL_510674\|2 | ···················· | ···················· | ···················· | ···················· | ···················· | ···················· |
| F | 980 hCoV-19/South Korea/KCDC2339/2020\|EPI_ISL_510675\|2 | ···················· | ···················· | ···················· | ···················· | ···················· | ···················· |
| F | 981 hCoV-19/South Korea/KCDC2340/2020\|EPI_ISL_510676\|2 | ···················· | ···················· | ···················· | ···················· | ···················· | ···················· |
| F | 982 hCoV-19/South Korea/KCDC2341/2020\|EPI_ISL_510677\|2 | ···················· | ···················· | ···················· | ···················· | ···················· | ···················· |
| F | 983 hCoV-19/South Korea/KCDC2342/2020\|EPI_ISL_510678\|2 | ···················· | ···················· | ···················· | ···················· | ···················· | ···················· |
| F | 984 hCoV-19/South Korea/KCDC2343/2020\|EPI_ISL_510679\|2 | ···················· | ···················· | ···················· | ···················· | ···················· | ···················· |
| F | 985 hCoV-19/South Korea/KCDC2344/2020\|EPI_ISL_510680\|2 | ···················· | ···················· | ···················· | ···················· | ···················· | ···················· |
| F | 986 hCoV-19/South Korea/KCDC2345/2020\|EPI_ISL_510681\|2 | ···················· | ···················· | ···················· | ···················· | ···················· | ···················· |
| F | 987 hCoV-19/South Korea/KCDC2346/2020\|EPI_ISL_510682\|2 | ···················· | ···················· | ···················· | ···················· | ···················· | ···················· |
| F | 988 hCoV-19/South Korea/KCDC2347/2020\|EPI_ISL_510683\|2 | ···················· | ···················· | ···················· | ···················· | ···················· | ···················· |
| F | 989 hCoV-19/South Korea/KCDC2348/2020\|EPI_ISL_510684\|2 | ···················· | ···················· | ···················· | ···················· | ···················· | ···················· |
| F | 990 hCoV-19/South Korea/KCDC2349/2020\|EPI_ISL_510685\|2 | ···················· | ···················· | ···················· | ···················· | ···················· | ···················· |
| F | 991 hCoV-19/South Korea/KCDC2350/2020\|EPI_ISL_510686\|2 | ···················· | ···················· | ···················· | ···················· | ···················· | ···················· |
| F | 992 hCoV-19/South Korea/KCDC2351/2020\|EPI_ISL_510687\|2 | ···················· | ···················· | ···················· | ···················· | ···················· | ···················· |
| F | 993 hCoV-19/South Korea/KCDC2352/2020\|EPI_ISL_510688\|2 | ···················· | ···················· | ···················· | ···················· | ···················· | ···················· |
| F | 994 hCoV-19/Spain/CT-IrsiCaixa-2020030095/2020\|EPI_ISL | ···················· | ···················· | ···················· | ···················· | ···················· | ···················· |
| F | 995 hCoV-19/Switzerland/BL-ETHZ-200000/2020\|EPI_ISL_51 | ···················· | ···················· | ···················· | ···················· | ···················· | ···················· |
| F | 996 hCoV-19/Switzerland/BL-ETHZ-200001/2020\|EPI_ISL_51 | ···················· | ···················· | ···················· | ···················· | ···················· | ···················· |
| F | 997 hCoV-19/Switzerland/AG-ETHZ-200002/2020\|EPI_ISL_51 | ···················· | ···················· | ···················· | ···················· | ···················· | ···················· |
| F | 998 hCoV-19/Switzerland/AG-ETHZ-200003/2020\|EPI_ISL_51 | ···················· | ···················· | ···················· | ···················· | ···················· | ···················· |
| F | 999 hCoV-19/Switzerland/BL-ETHZ-200004/2020\|EPI_ISL_51 | ···················· | ···················· | ···················· | ···················· | ···················· | ···················· |
| F | 1000 hCoV-19/Switzerland/BE-ETHZ-200005/2020\|EPI_ISL_51 | ···················· | ···················· | ···················· | ···················· | ···················· | ···················· |
| F | 1001 hCoV-19/Switzerland/BL-ETHZ-200006/2020\|EPI_ISL_51 | ···················· | ···················· | ···················· | ···················· | ···················· | ···················· |
| F | 1002 hCoV-19/Switzerland/GR-ETHZ-200009/2020\|EPI_ISL_51 | ···················· | ···················· | ···················· | ···················· | ···················· | ···················· |
| F | 1003 hCoV-19/Switzerland/VD-ETHZ-200010/2020\|EPI_ISL_51 | ···················· | ···················· | ···················· | ···················· | ···················· | ···················· |
| F | 1004 hCoV-19/Switzerland/SO-ETHZ-210070/2020\|EPI_ISL_51 | ···················· | ···················· | ···················· | ···················· | ···················· | ···················· |
| F | 1005 hCoV-19/Switzerland/BL-ETHZ-210074/2020\|EPI_ISL_51 | ···················· | ···················· | ···················· | ···················· | ·············Y······ | ···················· |
| F | 1006 hCoV-19/Switzerland/GR-ETHZ-210075/2020\|EPI_ISL_51 | ···········T········ | ···················· | ···················· | ···················· | ···················· | ···················· |
| F | 1007 hCoV-19/Switzerland/GR-ETHZ-210078/2020\|EPI_ISL_51 | ···········T········ | ···················· | ···················· | ···················· | ···················· | ···················· |
| F | 1008 hCoV-19/Switzerland/GR-ETHZ-210082/2020\|EPI_ISL_51 | ···········T········ | ···················· | ···················· | ···················· | ···················· | ···················· |
| F | 1009 hCoV-19/Switzerland/GR-ETHZ-210083/2020\|EPI_ISL_51 | ···········T········ | ···················· | ···················· | ···················· | ···················· | ···················· |
| F | 1010 hCoV-19/Switzerland/GE-ETHZ-210084/2020\|EPI_ISL_51 | ···················· | ···················· | ···················· | ···················· | ···················· | ···················· |
| F | 1011 hCoV-19/Switzerland/ZH-ETHZ-220010/2020\|EPI_ISL_51 | ···················· | ···················· | ···················· | ···················· | ···················· | ···················· |
| F | 1012 hCoV-19/Switzerland/BL-ETHZ-220011/2020\|EPI_ISL_51 | ···················· | ···················· | ···················· | ···················· | ···················· | ···················· |
| F | 1013 hCoV-19/Switzerland/AG-ETHZ-220012/2020\|EPI_ISL_51 | ···················· | ···················· | ··Y················· | ···················· | ···················· | ···················· |
| F | 1014 hCoV-19/SaudiArabia/KAUST-MADINAH1049/2020\|EPI_ISL | ···················· | ·A·················· | ···················· | ···················· | ···················· | ···A················ |
| F | 1015 hCoV-19/SaudiArabia/KAUST-MADINAH1051/2020\|EPI_ISL | ···················· | ·A·················· | ···················· | ···················· | ···················· | ···A················ |
| F | 1016 hCoV-19/SaudiArabia/KAUST-MADINAH1052/2020\|EPI_ISL | ···················· | ···················· | ···················· | ···················· | ···················· | ···················· |
| F | 1017 hCoV-19/SaudiArabia/KAUST-MADINAH1053/2020\|EPI_ISL | ···················· | ·A·················· | ···················· | ···················· | ···················· | ···A················ |
| F | 1018 hCoV-19/SaudiArabia/KAUST-MADINAH1060/2020\|EPI_ISL | ···················· | ·A·················· | ···················· | ···················· | ···················· | ···A················ |
| F | 1019 hCoV-19/SaudiArabia/KAUST-MADINAH1061/2020\|EPI_ISL | ···················· | ···················· | ···················· | ···················· | ···················· | ···················· |
| F | 1020 hCoV-19/SaudiArabia/KAUST-MADINAH1066/2020\|EPI_ISL | ···················· | ···················· | ···················· | ···················· | ···················· | ···················· |
| F | 1021 hCoV-19/SaudiArabia/KAUST-MADINAH1072/2020\|EPI_ISL | ···················· | ···················· | ···················· | ···················· | ···················· | ···················· |
| F | 1022 hCoV-19/SaudiArabia/KAUST-MADINAH1075/2020\|EPI_ISL | ···················· | ·A·················· | ···················· | ···················· | ···················· | ···A················ |
| F | 1023 hCoV-19/SaudiArabia/KAUST-MADINAH1077/2020\|EPI_ISL | ···················· | ·A·················· | ···················· | ···················· | ···················· | ···A················ |

| | | | | | | | | |
|---|---|---|---|---|---|---|---|---|
| F | 1024 | hCoV-19/SaudiArabia/KAUST-MADINAH1078/2020\|EPI_ISL | ···················· | ·A·················· | ···················· | ···················· | ···················· | ···A················ |
| F | 1025 | hCoV-19/USA/WA-S1574/2020\|EPI_ISL_496000\|2020-05-2 | ···················· | ···················· | ···················· | ···················· | ···················· | ···················· |
| F | 1026 | hCoV-19/USA/WA-S1575/2020\|EPI_ISL_496001\|2020-05-2 | ···················· | ···················· | ···················· | ···················· | ···················· | ···················· |
| F | 1027 | hCoV-19/USA/WA-S1576/2020\|EPI_ISL_496002\|2020-05-2 | ···················· | ···················· | ···················· | ···················· | ···················· | ···················· |
| F | 1028 | hCoV-19/USA/WA-S1577/2020\|EPI_ISL_496003\|2020-05-2 | ···················· | ···················· | ···················· | ···················· | ···················· | ···················· |
| F | 1029 | hCoV-19/USA/WA-S1578/2020\|EPI_ISL_496004\|2020-05-2 | ···················· | ···················· | ···················· | ···················· | ···················· | ···················· |
| F | 1030 | hCoV-19/USA/WA-S1579/2020\|EPI_ISL_496005\|2020-05-2 | ···················· | ···················· | ···················· | ···················· | ···················· | ···················· |
| F | 1031 | hCoV-19/USA/WA-S1580/2020\|EPI_ISL_496006\|2020-05-2 | ···················· | ···················· | ···················· | ···················· | ···················· | ···················· |
| F | 1032 | hCoV-19/USA/WA-S1581/2020\|EPI_ISL_496007\|2020-05-2 | ···················· | ·A·················· | ···················· | ···················· | ···················· | ···················· |
| F | 1033 | hCoV-19/USA/WA-S1582/2020\|EPI_ISL_496008\|2020-05-2 | ···················· | ···················· | ···················· | ···················· | ···················· | ···················· |
| F | 1034 | hCoV-19/USA/WA-S1583/2020\|EPI_ISL_496009\|2020-05-2 | ···················· | ···················· | ···················· | ···················· | ···················· | ···················· |
| F | 1035 | hCoV-19/USA/WA-S1585/2020\|EPI_ISL_496011\|2020-05-2 | ···················· | ···················· | ···················· | ···················· | ···················· | ···················· |
| F | 1036 | hCoV-19/USA/WA-S1586/2020\|EPI_ISL_496012\|2020-05-2 | ···················· | ···················· | ···················· | ···················· | ···················· | ···················· |
| F | 1037 | hCoV-19/USA/WA-S1587/2020\|EPI_ISL_496013\|2020-05-2 | ···················· | ···················· | ···················· | ···················· | ···················· | ···················· |
| F | 1038 | hCoV-19/USA/WA-S1588/2020\|EPI_ISL_496014\|2020-05-2 | ···················· | ···················· | ···················· | ···················· | ···················· | ···················· |
| F | 1039 | hCoV-19/USA/WA-S1590/2020\|EPI_ISL_496016\|2020-05-2 | ···················· | ···················· | ···················· | ···················· | ···················· | ···················· |
| F | 1040 | hCoV-19/USA/WA-S1592/2020\|EPI_ISL_496018\|2020-05-2 | ···················· | ···················· | ···················· | ···················· | ···················· | ···················· |
| F | 1041 | hCoV-19/USA/WA-S1593/2020\|EPI_ISL_496019\|2020-05-2 | ···················· | ···················· | ···················· | ···················· | ···················· | ···················· |
| F | 1042 | hCoV-19/USA/WA-S1595/2020\|EPI_ISL_496021\|2020-05-2 | ···················· | ···················· | ···················· | ···················· | ···················· | ···················· |
| F | 1043 | hCoV-19/USA/WA-S1596/2020\|EPI_ISL_496022\|2020-05-2 | ···················· | ···················· | ···················· | ···················· | ···················· | ···················· |
| F | 1044 | hCoV-19/USA/WA-S1597/2020\|EPI_ISL_496023\|2020-05-2 | ···················· | ···················· | ···················· | ···················· | ···················· | ···················· |
| F | 1045 | hCoV-19/USA/WA-S1598/2020\|EPI_ISL_496024\|2020-05-2 | ···················· | ···················· | ···················· | ···················· | ···················· | ···················· |
| F | 1046 | hCoV-19/USA/WA-S1599/2020\|EPI_ISL_496025\|2020-05-2 | ···················· | ···················· | ···················· | ···················· | ···················· | ···················· |
| F | 1047 | hCoV-19/USA/WA-S1600/2020\|EPI_ISL_496026\|2020-05-2 | ···················· | ···················· | ···················· | ···················· | ···················· | ···················· |
| F | 1048 | hCoV-19/USA/WA-S1601/2020\|EPI_ISL_496027\|2020-05-2 | ···················· | ···················· | ···················· | ···················· | ···················· | ···················· |
| F | 1049 | hCoV-19/USA/WA-S1602/2020\|EPI_ISL_496028\|2020-05-2 | ···················· | ···················· | ···················· | ···················· | ···················· | ···················· |
| F | 1050 | hCoV-19/USA/WA-S1603/2020\|EPI_ISL_496029\|2020-05-2 | ···················· | ···················· | ···················· | ···················· | ···················· | ···················· |
| F | 1051 | hCoV-19/USA/WA-S1604/2020\|EPI_ISL_496030\|2020-05-2 | ···················· | ···················· | ···················· | ···················· | ···················· | ···················· |
| F | 1052 | hCoV-19/USA/WA-S1605/2020\|EPI_ISL_496031\|2020-05-2 | ···················· | ···················· | ···················· | ···················· | ···················· | ···················· |
| F | 1053 | hCoV-19/USA/WA-S1606/2020\|EPI_ISL_496032\|2020-05-2 | ···················· | ···················· | ···················· | ···················· | ···················· | ···················· |
| F | 1054 | hCoV-19/USA/WA-S1608/2020\|EPI_ISL_496034\|2020-05-2 | ···················· | ···················· | ···················· | ···················· | ···················· | ···················· |
| F | 1055 | hCoV-19/USA/WA-S1609/2020\|EPI_ISL_496035\|2020-05-2 | ···················· | ···················· | ···················· | ···················· | ···················· | ···················· |
| F | 1056 | hCoV-19/USA/WA-S1617/2020\|EPI_ISL_496043\|2020-05-2 | ···················· | ···················· | ···················· | ···················· | ···················· | ···················· |
| F | 1057 | hCoV-19/USA/WA-S1618/2020\|EPI_ISL_496044\|2020-05-2 | ···················· | ···················· | ···················· | ···················· | ···················· | ···················· |
| F | 1058 | hCoV-19/USA/WA-S1632/2020\|EPI_ISL_496058\|2020-05-2 | ···················· | ···················· | ···················· | ···················· | ···················· | ···················· |
| F | 1059 | hCoV-19/USA/WA-S1633/2020\|EPI_ISL_496059\|2020-05-2 | ···················· | ···················· | ···················· | ···················· | ···················· | ···················· |
| F | 1060 | hCoV-19/USA/WA-S1634/2020\|EPI_ISL_496060\|2020-05-2 | ···················· | ···················· | ···················· | ···················· | ···················· | ···················· |
| F | 1061 | hCoV-19/USA/WA-S1635/2020\|EPI_ISL_496061\|2020-05-2 | ···················· | ···················· | ···················· | ···················· | ···················· | ···················· |
| F | 1062 | hCoV-19/USA/WA-S1636/2020\|EPI_ISL_496062\|2020-05-2 | ···················· | ···················· | ···················· | ···················· | ···················· | ···················· |
| F | 1063 | hCoV-19/USA/WA-S1637/2020\|EPI_ISL_496063\|2020-05-2 | ···················· | ···················· | ···················· | ···················· | ···················· | ···················· |
| F | 1064 | hCoV-19/USA/WA-S1638/2020\|EPI_ISL_496064\|2020-05-2 | ···················· | ···················· | ···················· | ···················· | ···················· | ···················· |
| F | 1065 | hCoV-19/USA/WA-S1639/2020\|EPI_ISL_496065\|2020-05-2 | ···················· | ···················· | ···················· | ···················· | ···················· | ···················· |
| F | 1066 | hCoV-19/USA/WA-S1640/2020\|EPI_ISL_496066\|2020-05-2 | ···················· | ···················· | ···················· | ···················· | ···················· | ···················· |
| F | 1067 | hCoV-19/USA/WA-S1641/2020\|EPI_ISL_496067\|2020-05-2 | ···················· | ···················· | ···················· | ···················· | ···················· | ···················· |
| F | 1068 | hCoV-19/USA/WA-S1642/2020\|EPI_ISL_496068\|2020-05-2 | ···················· | ···················· | ···················· | ···················· | ···················· | ···················· |
| F | 1069 | hCoV-19/USA/WA-S1643/2020\|EPI_ISL_496069\|2020-05-2 | ···················· | ···················· | ···················· | ···················· | ···················· | ···················· |
| F | 1070 | hCoV-19/USA/WA-S1644/2020\|EPI_ISL_496070\|2020-05-2 | ···················· | ···················· | ···················· | ···················· | ···················· | ···················· |
| F | 1071 | hCoV-19/USA/WA-S1645/2020\|EPI_ISL_496071\|2020-05-2 | ···················· | ···················· | ···················· | ···················· | ···················· | ···················· |
| F | 1072 | hCoV-19/USA/WA-S1647/2020\|EPI_ISL_496073\|2020-05-2 | ···················· | ···················· | ···················· | ···················· | ···················· | ···················· |
| F | 1073 | hCoV-19/USA/WA-S1648/2020\|EPI_ISL_496074\|2020-05-2 | ···················· | ···················· | ···················· | ···················· | ···················· | ···················· |
| F | 1074 | hCoV-19/USA/WA-S1650/2020\|EPI_ISL_496076\|2020-05-2 | ···················· | ···················· | ···················· | ···················· | ···················· | ···················· |
| F | 1075 | hCoV-19/USA/WA-S1651/2020\|EPI_ISL_496077\|2020-05-2 | ···················· | ···················· | ···················· | ···················· | ···················· | ···················· |
| F | 1076 | hCoV-19/USA/WA-S1652/2020\|EPI_ISL_496078\|2020-05-2 | ···················· | ···················· | ···················· | ···················· | ···················· | ···················· |
| F | 1077 | hCoV-19/USA/WA-S1653/2020\|EPI_ISL_496079\|2020-05-2 | ···················· | ···················· | ···················· | ···················· | ···················· | ···················· |
| F | 1078 | hCoV-19/USA/WA-S2691/2020\|EPI_ISL_497323\|2020-06-2 | ···················· | ···················· | ···················· | ···················· | ···················· | ···················· |
| F | 1079 | hCoV-19/USA/WA-S2692/2020\|EPI_ISL_497324\|2020-06-2 | ···················· | ···················· | ···················· | ···················· | ···················· | ···················· |
| F | 1080 | hCoV-19/USA/WA-S2693/2020\|EPI_ISL_497325\|2020-06-2 | ···················· | ···················· | ···················· | ···················· | ···················· | ···················· |
| F | 1081 | hCoV-19/USA/WA-S2694/2020\|EPI_ISL_497326\|2020-06-2 | ···················· | ···················· | ···················· | ···················· | ···················· | ···················· |
| F | 1082 | hCoV-19/USA/WA-S2695/2020\|EPI_ISL_497327\|2020-06-2 | ···················· | ···················· | ···················· | ···················· | ···················· | ···················· |
| F | 1083 | hCoV-19/USA/WA-S2697/2020\|EPI_ISL_497329\|2020-06-2 | ···················· | ···················· | ···················· | ···················· | ···················· | ···················· |
| F | 1084 | hCoV-19/USA/WA-S2698/2020\|EPI_ISL_497330\|2020-06-2 | ···················· | ···················· | ···················· | ···················· | ···················· | ···················· |
| F | 1085 | hCoV-19/USA/WA-S2699/2020\|EPI_ISL_497331\|2020-06-2 | ···················· | ···················· | ···················· | ···················· | ···················· | ···················· |
| F | 1086 | hCoV-19/USA/WA-S2700/2020\|EPI_ISL_497332\|2020-06-2 | ···················· | ···················· | ···················· | ···················· | ···················· | ···················· |
| F | 1087 | hCoV-19/USA/WA-S2701/2020\|EPI_ISL_497333\|2020-06-2 | ···················· | ···················· | ···················· | ···················· | ···················· | ···················· |
| F | 1088 | hCoV-19/USA/WA-S2703/2020\|EPI_ISL_497335\|2020-06-2 | ···················· | ···················· | ···················· | ···················· | ···················· | ···················· |
| F | 1089 | hCoV-19/USA/WA-S2704/2020\|EPI_ISL_497336\|2020-06-2 | ···················· | ···················· | ···················· | ···················· | ···················· | ···················· |
| F | 1090 | hCoV-19/USA/WA-S2705/2020\|EPI_ISL_497337\|2020-06-2 | ···················· | ···················· | ···················· | ···················· | ···················· | ···················· |
| F | 1091 | hCoV-19/USA/WA-S2706/2020\|EPI_ISL_497338\|2020-06-2 | ···················· | ···················· | ···················· | ···················· | ···················· | ···················· |
| F | 1092 | hCoV-19/USA/WA-S2707/2020\|EPI_ISL_497339\|2020-06-2 | ···················· | ···················· | ···················· | ···················· | ···················· | ···················· |
| F | 1093 | hCoV-19/USA/WA-S2708/2020\|EPI_ISL_497340\|2020-06-2 | ···················· | ···················· | ···················· | ···················· | ···················· | ···················· |
| F | 1094 | hCoV-19/USA/CA-S1920/2020\|EPI_ISL_497341\|2020-06-0 | ···················· | ···················· | ···················· | ···················· | ···················· | ···················· |
| F | 1095 | hCoV-19/USA/WA-S1913/2020\|EPI_ISL_497342\|2020-06-0 | ···················· | ···················· | ···················· | ···················· | ···················· | ···················· |
| F | 1096 | hCoV-19/USA/WA-S1914/2020\|EPI_ISL_497343\|2020-06-0 | ···················· | ···················· | ···················· | ···················· | ···················· | ···················· |
| F | 1097 | hCoV-19/USA/WA-S1915/2020\|EPI_ISL_497344\|2020-06-0 | ···················· | ···················· | ···················· | ···················· | ···················· | ···················· |
| F | 1098 | hCoV-19/USA/WA-S1916/2020\|EPI_ISL_497345\|2020-06-0 | ···················· | ···················· | ···················· | ···················· | ···················· | ···················· |
| F | 1099 | hCoV-19/USA/WA-S1917/2020\|EPI_ISL_497346\|2020-06-0 | ···················· | ···················· | ···················· | ···················· | ···················· | ···················· |
| F | 1100 | hCoV-19/USA/WA-S1918/2020\|EPI_ISL_497347\|2020-06-0 | ···················· | ···················· | ···················· | ···················· | ···················· | ···················· |
| F | 1101 | hCoV-19/USA/WA-S1919/2020\|EPI_ISL_497348\|2020-06-0 | ···················· | ···················· | ···················· | ···················· | ···················· | ···················· |
| F | 1102 | hCoV-19/USA/WA-S1921/2020\|EPI_ISL_497349\|2020-06-0 | ···················· | ···················· | ···················· | ···················· | ···················· | ···················· |
| F | 1103 | hCoV-19/USA/WA-S1922/2020\|EPI_ISL_497350\|2020-06-0 | ···················· | ···················· | ···················· | ···················· | ···················· | ···················· |

F 1104 hCoV-19/USA/WA-S1923/2020|EPI_ISL_497351|2020-06-0 ···················· ···················· ····················· ···················· ···················· ····················
F 1105 hCoV-19/USA/WA-S1924/2020|EPI_ISL_497352|2020-06-0 ···················· ···················· ····················· ···················· ···················· ····················
F 1106 hCoV-19/USA/WA-S1925/2020|EPI_ISL_497353|2020-06-0 ···················· ···················· ····················· ···················· ···················· ····················
F 1107 hCoV-19/USA/WA-S1926/2020|EPI_ISL_497354|2020-06-0 ···················· ···················· ····················· ···················· ···················· ····················
F 1108 hCoV-19/USA/WA-S1928/2020|EPI_ISL_497356|2020-06-0 ···················· ···················· ····················· ···················· ···················· ····················
F 1109 hCoV-19/USA/WA-S1929/2020|EPI_ISL_497357|2020-06-0 ···················· ···················· ····················· ···················· ···················· ····················
F 1110 hCoV-19/USA/WA-S1930/2020|EPI_ISL_497358|2020-06-0 ···················· ···················· ····················· ···················· ···················· ····················
F 1111 hCoV-19/USA/WA-S1931/2020|EPI_ISL_497359|2020-06-0 ···················· ···················· ····················· ···················· ···················· ····················
F 1112 hCoV-19/USA/WA-S1932/2020|EPI_ISL_497360|2020-06-0 ···················· ···················· ····················· ···················· ···················· ····················
F 1113 hCoV-19/USA/WA-S1933/2020|EPI_ISL_497361|2020-06-0 ···················· ···················· ····················· ···················· ···················· ····················
F 1114 hCoV-19/USA/WA-S1934/2020|EPI_ISL_497362|2020-06-0 ···················· ···················· ····················· ···················· ···················· ····················
F 1115 hCoV-19/USA/WA-S1935/2020|EPI_ISL_497363|2020-06-0 ···················· ···················· ····················· ···················· ···················· ····················
F 1116 hCoV-19/USA/WA-S1936/2020|EPI_ISL_497364|2020-06-0 ···················· ···················· ····················· ···················· ···················· ····················
F 1117 hCoV-19/USA/WA-S1937/2020|EPI_ISL_497365|2020-06-0 ···················· ···················· ····················· ···················· ···················· ····················
F 1118 hCoV-19/USA/WA-S1938/2020|EPI_ISL_497366|2020-06-0 ···················· ···················· ····················· ···················· ···················· ····················
F 1119 hCoV-19/USA/WA-S1946/2020|EPI_ISL_497374|2020-06-0 ···················· ···················· ····················· ···················· ···················· ····················
F 1120 hCoV-19/USA/WA-S1947/2020|EPI_ISL_497375|2020-06-0 ···················· ···················· ····················· ···················· ···················· ····················
F 1121 hCoV-19/USA/WA-S1961/2020|EPI_ISL_497389|2020-06-0 ···················· ···················· ····················· ···················· ···················· ····················
F 1122 hCoV-19/USA/WA-S1962/2020|EPI_ISL_497390|2020-06-0 ···················· ···················· ····················· ···················· ···················· ····················
F 1123 hCoV-19/USA/WA-S1963/2020|EPI_ISL_497391|2020-06-0 ···················· ···················· ····················· ···················· ···················· ····················
F 1124 hCoV-19/USA/WA-S1964/2020|EPI_ISL_497392|2020-06-0 ···················· ···················· ····················· ···················· ···················· ····················
F 1125 hCoV-19/USA/WA-S1965/2020|EPI_ISL_497393|2020-06-0 ···················· ···················· ····················· ···················· ···················· ····················
F 1126 hCoV-19/USA/WA-S1966/2020|EPI_ISL_497394|2020-06-0 ···················· ···················· ····················· ···················· ···················· ····················
F 1127 hCoV-19/USA/WA-S1967/2020|EPI_ISL_497395|2020-06-0 ···················· ···················· ····················· ···················· ···················· ····················
F 1128 hCoV-19/USA/WA-S1968/2020|EPI_ISL_497396|2020-06-0 ···················· ···················· ····················· ···················· ···················· ····················
F 1129 hCoV-19/USA/WA-S1969/2020|EPI_ISL_497397|2020-06-0 ···················· ···················· ····················· ···················· ···················· ····················
F 1130 hCoV-19/USA/WA-S1970/2020|EPI_ISL_497398|2020-06-0 ···················· ····A··············· ····················· ···················· ···················· ····················
F 1131 hCoV-19/USA/WA-S1971/2020|EPI_ISL_497399|2020-06-0 ···················· ···················· ····················· ···················· ···················· ····················
F 1132 hCoV-19/Switzerland/ZH-ETHZ-190005/2020|EPI_ISL_49 ···················· ·A·················· ····················· ···················· ···················· ····················
F 1133 hCoV-19/Switzerland/BL-ETHZ-190006/2020|EPI_ISL_49 ···················· ···················· ····················· ···················· ···················· ····················
F 1134 hCoV-19/USA/UT-03372/2020|EPI_ISL_498654|2020-04-2 ···················· ···················· ····················· ···················· ···················· ····················
F 1135 hCoV-19/USA/UT-03378/2020|EPI_ISL_498655|2020-04-2 ···················· ···················· ····················· ···················· ···················· ····················
F 1136 hCoV-19/USA/UT-03379/2020|EPI_ISL_498656|2020-04-2 ···················· ···················· ····················· ···················· ···················· ····················
F 1137 hCoV-19/USA/UT-03380/2020|EPI_ISL_498657|2020-04-2 ···················· ···················· ····················· ···················· ···················· ····················
F 1138 hCoV-19/USA/UT-03428/2020|EPI_ISL_498658|2020-04-2 ···················· ···················· ····················· ···················· ···················· ····················
F 1139 hCoV-19/USA/UT-03429/2020|EPI_ISL_498659|2020-04-2 ···················· ···················· ····················· ···················· ···················· ····················
F 1140 hCoV-19/USA/UT-03431/2020|EPI_ISL_498660|2020-04-2 ···················· ···················· ····················· ···················· ···················· ····················
F 1141 hCoV-19/USA/UT-03432/2020|EPI_ISL_498661|2020-04-2 ···················· ···················· ····················· ···················· ···················· ····················
F 1142 hCoV-19/USA/UT-03433/2020|EPI_ISL_498662|2020-04-2 ···················· ···················· ····················· ···················· ···················· ····················
F 1143 hCoV-19/USA/UT-03434/2020|EPI_ISL_498663|2020-04-2 ···················· ···················· ····················· ···················· ···················· ····················
F 1144 hCoV-19/USA/UT-03435/2020|EPI_ISL_498664|2020-04-2 ···················· ···················· ····················· ···················· ···················· ····················
F 1145 hCoV-19/USA/UT-03436/2020|EPI_ISL_498665|2020-04-2 ···················· ···················· ····················· ···················· ···················· ····················
F 1146 hCoV-19/USA/UT-03437/2020|EPI_ISL_498666|2020-04-2 ···················· ···················· ····················· ···················· ···················· ····················
F 1147 hCoV-19/USA/UT-03438/2020|EPI_ISL_498667|2020-04-2 ···················· ···················· ····················· ···················· ···················· ····················
F 1148 hCoV-19/USA/UT-02898/2020|EPI_ISL_498668|2020-04-3 ···················· ···················· ····················· ···················· ···················· ····················
F 1149 hCoV-19/USA/UT-02938/2020|EPI_ISL_498670|2020-04-3 ···················· ···················· ····················· ···················· ···················· ····················
F 1150 hCoV-19/USA/UT-02939/2020|EPI_ISL_498671|2020-04-3 ···················· ···················· ····················· ···················· ···················· ····················
F 1151 hCoV-19/USA/UT-04209/2020|EPI_ISL_498687|2020-05-1 ···················· ···················· ····················· ···················· ···················· ····················
F 1152 hCoV-19/Liaoning/IVDC-04/2020|EPI_ISL_498691|2020- ···················· ···················· ····················· ···················· ···················· ····················
F 1153 hCoV-19/Liaoning/IVDC-01/2020|EPI_ISL_498692|2020- ···················· ···················· ····················· ···················· ···················· ····················
F 1154 hCoV-19/Liaoning/IVDC-03/2020|EPI_ISL_498693|2020- ···················· ···················· ····················· ···················· ···················· ····················
F 1155 hCoV-19/Liaoning/IVDC-02/2020|EPI_ISL_498694|2020- ···················· ···················· ····················· ···················· ···················· ····················
F 1156 hCoV-19/USA/AL-QDX-244/2020|EPI_ISL_498695|2020-04 ···················· ···················· ····················· ···················· ···················· ····················
F 1157 hCoV-19/USA/AL-QDX-245/2020|EPI_ISL_498696|2020-04 ···················· ···················· ····················· ···················· ···················· ····················
F 1158 hCoV-19/USA/CA-QDX-221/2020|EPI_ISL_498697|2020-04 ···················· ···················· ····················· ···················· ···················· ····················
F 1159 hCoV-19/England/LIVE-A1229/2020|EPI_ISL_499985|202 ···················· ·A·················· ····················· ···················· ···················· ····················
F 1160 hCoV-19/England/LIVE-A1247/2020|EPI_ISL_499986|202 ···················· ·A·················· ····················· ···················· ···················· ····················
F 1168 hCoV-19/England/LIVE-A1344/2020|EPI_ISL_499994|202 ···················· ·A·················· ····················· ···················· ···················· ····················
F 1169 hCoV-19/England/LIVE-A245F/2020|EPI_ISL_499995|202 ···················· ···················· ····················· ···················· ···················· ····················
F 1170 hCoV-19/England/LIVE-A254D/2020|EPI_ISL_499996|202 ···················· ···················· ····················· ···················· ···················· ····················
F 1171 hCoV-19/England/LIVE-A255C/2020|EPI_ISL_499997|202 ···················· ···················· ····················· ···················· ···················· ····················
F 1172 hCoV-19/England/LIVE-A256B/2020|EPI_ISL_499998|202 ···················· ···················· ····················· ···················· ···················· ····················
F 1173 hCoV-19/England/LIVE-A25D4/2020|EPI_ISL_499999|202 ···················· ···················· ····················· ···················· ···················· ····················
F 1174 hCoV-19/Singapore/31nan/2020|EPI_ISL_507002|2020-0 ···················· ···················· ····················· ···················· ···················· ····················
F 1175 hCoV-19/Singapore/758/2020|EPI_ISL_507003|2020-03- ···················· ···················· ····················· ···················· ···················· ····················
F 1176 hCoV-19/Singapore/759/2020|EPI_ISL_507004|2020-03- ···················· ···················· ····················· ···················· ···················· ····················
F 1177 hCoV-19/Singapore/760/2020|EPI_ISL_507005|2020-04- ···················· ···················· ····················· ···················· ···················· ····················
F 1178 hCoV-19/Singapore/761/2020|EPI_ISL_507006|2020-04- ···················· ···················· ····················· ···················· ···················· ····················
F 1179 hCoV-19/Sweden/SVA200327SZ-NSW/2020|EPI_ISL_507008 ···················· ···················· ····················· ···················· ···················· ····················
F 1180 hCoV-19/Saudi Arabia/KAIMRC01/2020|EPI_ISL_507010| ···················· ···················· ····················· ···················· ···················· ····················
F 1181 hCoV-19/Saudi Arabia/KAIMRC02/2020|EPI_ISL_507011| ···················· ···················· ····················· ···················· ···················· ····················
F 1182 hCoV-19/Saudi Arabia/KAIMRC03/2020|EPI_ISL_507012| ···················· ···················· ····················· ···················· ···················· ····················
F 1183 hCoV-19/Saudi Arabia/KAIMRC07/2020|EPI_ISL_507013| ···················· ···················· ····················· ···················· ···················· ····················
F 1184 hCoV-19/Saudi Arabia/KAIMRC08/2020|EPI_ISL_507014| ···················· ···················· ····················· ···················· ···················· ····················
F 1185 hCoV-19/Saudi Arabia/KAIMRC09/2020|EPI_ISL_507015| ···················· ···················· ····················· ···················· ···················· ····················
F 1186 hCoV-19/Saudi Arabia/KAIMRC10/2020|EPI_ISL_507016| ···················· ···················· ····················· ···················· ···················· ····················
F 1187 hCoV-19/Saudi Arabia/KAIMRC11/2020|EPI_ISL_507017| ···················· ···················· ····················· ···················· ···················· ····················
F 1188 hCoV-19/Saudi Arabia/KAIMRC12/2020|EPI_ISL_507018| ···················· ···················· ····················· ···················· ···················· ····················
F 1189 hCoV-19/Saudi Arabia/KAIMRC13/2020|EPI_ISL_507019| ···················· ···················· ····················· ···················· ···················· ····················
F 1190 hCoV-19/India/KA-IB53/2020|EPI_ISL_508330|2020-06- ···················· ···················· ····················· ···················· ···················· ····················

| | | | | | | | |
|---|---|---|---|---|---|---|---|
| F | 1191 hCoV-19/India/KA-IB54/2020\|EPI_ISL_508331\|2020-06- | ···················· | ···················· | ···················· | ···················· | ···················· | ···················· |
| F | 1192 hCoV-19/India/KA-IB58/2020\|EPI_ISL_508332\|2020-05- | ···················· | ·A·················· | ···················· | ···················· | ···················· | ···A················ |
| F | 1193 hCoV-19/India/KA-IB6/2020\|EPI_ISL_508333\|2020-06-1 | ···················· | ···················· | ···················· | ···················· | ···················· | ···················· |
| F | 1194 hCoV-19/India/KA-IB62/2020\|EPI_ISL_508334\|2020-05- | ···················· | ···················· | ···················· | ···················· | ···················· | ···················· |
| F | 1195 hCoV-19/India/KA-IB7/2020\|EPI_ISL_508335\|2020-06-1 | ···················· | ···················· | ···················· | ···················· | ···················· | ···················· |
| F | 1196 hCoV-19/India/KA-IB8/2020\|EPI_ISL_508336\|2020-06-2 | ···················· | ···················· | ···················· | ···················· | ···················· | ···················· |
| F | 1197 hCoV-19/India/KA-IB9/2020\|EPI_ISL_508337\|2020-06-1 | ···················· | ···················· | ···················· | ···················· | ···················· | ···················· |
| F | 1198 hCoV-19/India/WB-IK1/2020\|EPI_ISL_508338\|2020-03-3 | ···················· | ···················· | ···················· | ···················· | ···················· | ···················· |
| F | 1199 hCoV-19/India/WB-IK10/2020\|EPI_ISL_508339\|2020-04- | ···················· | ···················· | ···················· | ···················· | ···A················ | ···················· |
| F | 1200 hCoV-19/India/WB-IK11/2020\|EPI_ISL_508340\|2020-04- | ···················· | ···················· | ···················· | ···················· | ···················· | ···················· |
| F | 1201 hCoV-19/India/WB-IK12/2020\|EPI_ISL_508341\|2020-04- | ···················· | ···················· | ···················· | ···················· | ···A················ | ···················· |
| F | 1202 hCoV-19/India/WB-IK13/2020\|EPI_ISL_508342\|2020-04- | ···················· | ···················· | ···················· | ···················· | ···················· | ···················· |
| F | 1203 hCoV-19/India/WB-IK14/2020\|EPI_ISL_508343\|2020-04- | ···················· | ···················· | ···················· | ···················· | ···················· | ···················· |
| F | 1204 hCoV-19/India/WB-IK15/2020\|EPI_ISL_508344\|2020-04- | ···················· | ···················· | ···················· | ···················· | ···················· | ···················· |
| F | 1205 hCoV-19/India/WB-IK16/2020\|EPI_ISL_508345\|2020-04- | ···················· | ···················· | ···················· | ···················· | ···················· | ···················· |
| F | 1206 hCoV-19/India/WB-IK17/2020\|EPI_ISL_508346\|2020-04- | ···················· | ···················· | ···················· | ···················· | ···················· | ···················· |
| F | 1207 hCoV-19/India/WB-IK18/2020\|EPI_ISL_508347\|2020-04- | ···················· | ···················· | ···················· | ···················· | ···················· | ···················· |
| F | 1208 hCoV-19/India/WB-IK19/2020\|EPI_ISL_508348\|2020-04- | ···················· | ···················· | ···················· | ···················· | ···················· | ···················· |
| F | 1209 hCoV-19/India/WB-IK2/2020\|EPI_ISL_508349\|2020-03-3 | ···················· | ···················· | ···················· | ···················· | ···················· | ···················· |
| F | 1210 hCoV-19/USA/GA-CDC-0851/2020\|EPI_ISL_509662\|2020-0 | ···················· | ·A·················· | ···················· | ···················· | ···················· | ···················· |
| F | 1211 hCoV-19/USA/AR-CDC-4169/2020\|EPI_ISL_509663\|2020-0 | ···················· | ···················· | ···················· | ···················· | ···················· | ···················· |
| F | 1212 hCoV-19/USA/AR-CDC-4201/2020\|EPI_ISL_509664\|2020-0 | ···················· | ···················· | ···················· | ···················· | ···················· | ···················· |
| F | 1213 hCoV-19/USA/AR-CDC-4170/2020\|EPI_ISL_509665\|2020-0 | ···················· | ···················· | ···················· | ···················· | ···················· | ···················· |
| F | 1214 hCoV-19/USA/AR-CDC-4202/2020\|EPI_ISL_509666\|2020-0 | ···················· | ···················· | ···················· | ···················· | ···················· | ···················· |
| F | 1215 hCoV-19/USA/AR-CDC-4174/2020\|EPI_ISL_509667\|2020-0 | ···················· | ···················· | ···················· | ···················· | ···················· | ···················· |
| F | 1216 hCoV-19/USA/AR-CDC-4198/2020\|EPI_ISL_509668\|2020-0 | ···················· | ···················· | ···················· | ···················· | ···················· | ···················· |
| F | 1217 hCoV-19/USA/AR-CDC-4178/2020\|EPI_ISL_509670\|2020-0 | ···················· | ···················· | ···················· | ···················· | ···················· | ···················· |
| F | 1218 hCoV-19/USA/AR-CDC-4186/2020\|EPI_ISL_509671\|2020-0 | ···················· | ···················· | ···················· | ···················· | ···················· | ···················· |
| F | 1219 hCoV-19/USA/AR-CDC-4187/2020\|EPI_ISL_509672\|2020-0 | ···················· | ···················· | ···················· | ···················· | ···················· | ···················· |
| F | 1220 hCoV-19/USA/AR-CDC-4190/2020\|EPI_ISL_509673\|2020-0 | ···················· | ···················· | ···················· | ···················· | ···················· | ···················· |
| F | 1221 hCoV-19/USA/AR-CDC-4191/2020\|EPI_ISL_509674\|2020-0 | ···················· | ···················· | ···················· | ···················· | ···················· | ···················· |
| F | 1222 hCoV-19/USA/AR-CDC-4185/2020\|EPI_ISL_509675\|2020-0 | ···················· | ···················· | ···················· | ···················· | ···················· | ···················· |
| F | 1223 hCoV-19/USA/AR-CDC-4193/2020\|EPI_ISL_509676\|2020-0 | ···················· | ···················· | ···················· | ···················· | ···················· | ···················· |
| F | 1224 hCoV-19/USA/AR-CDC-4196/2020\|EPI_ISL_509677\|2020-0 | ···················· | ···················· | ···················· | ···················· | ···················· | ···················· |
| F | 1225 hCoV-19/USA/AR-CDC-4195/2020\|EPI_ISL_509678\|2020-0 | ···················· | ···················· | ···················· | ···················· | ···················· | ···················· |
| F | 1226 hCoV-19/USA/AR-CDC-4171/2020\|EPI_ISL_509679\|2020-0 | ···················· | ···················· | ···················· | ···················· | ···················· | ···················· |
| F | 1227 hCoV-19/South Korea/KCDC2308/2020\|EPI_ISL_510644\|2 | ···················· | ···················· | ···················· | ···················· | ···················· | ···················· |
| F | 1228 hCoV-19/South Korea/KCDC2309/2020\|EPI_ISL_510645\|2 | ···················· | ···················· | ···················· | ···················· | ···················· | ···················· |
| F | 1229 hCoV-19/South Korea/KCDC2310/2020\|EPI_ISL_510646\|2 | ···················· | ···················· | ···················· | ···················· | ···················· | ···················· |
| F | 1230 hCoV-19/South Korea/KCDC2311/2020\|EPI_ISL_510647\|2 | ···················· | ···················· | ···················· | ···················· | ···················· | ···················· |
| F | 1231 hCoV-19/South Korea/KCDC2312/2020\|EPI_ISL_510648\|2 | ···················· | ···················· | ···················· | ···················· | ···················· | ···················· |
| F | 1232 hCoV-19/South Korea/KCDC2313/2020\|EPI_ISL_510649\|2 | ···················· | ···················· | ···················· | ···················· | ···················· | ···················· |
| F | 1233 hCoV-19/South Korea/KCDC2314/2020\|EPI_ISL_510650\|2 | ···················· | ···················· | ···················· | ···················· | ···················· | ···················· |
| F | 1234 hCoV-19/South Korea/KCDC2315/2020\|EPI_ISL_510651\|2 | ···················· | ···················· | ···················· | ···················· | ···················· | ···················· |
| F | 1235 hCoV-19/South Korea/KCDC2316/2020\|EPI_ISL_510652\|2 | ···················· | ···················· | ···················· | ···················· | ···················· | ···················· |
| F | 1236 hCoV-19/South Korea/KCDC2317/2020\|EPI_ISL_510653\|2 | ···················· | ···················· | ···················· | ···················· | ···················· | ···················· |
| F | 1237 hCoV-19/South Korea/KCDC2318/2020\|EPI_ISL_510654\|2 | ···················· | ···················· | ···················· | ···················· | ···················· | ···················· |
| F | 1238 hCoV-19/South Korea/KCDC2319/2020\|EPI_ISL_510655\|2 | ···················· | ···················· | ···················· | ···················· | ···················· | ···················· |
| F | 1239 hCoV-19/South Korea/KCDC2320/2020\|EPI_ISL_510656\|2 | ···················· | ···················· | ···················· | ···················· | ···················· | ···················· |
| F | 1240 hCoV-19/South Korea/KCDC2321/2020\|EPI_ISL_510657\|2 | ···················· | ···················· | ···················· | ···················· | ···················· | ···················· |
| F | 1241 hCoV-19/South Korea/KCDC2322/2020\|EPI_ISL_510658\|2 | ···················· | ···················· | ···················· | ···················· | ···················· | ···················· |
| F | 1242 hCoV-19/South Korea/KCDC2323/2020\|EPI_ISL_510659\|2 | ···················· | ···················· | ···················· | ···················· | ···················· | ···················· |
| F | 1243 hCoV-19/South Korea/KCDC2324/2020\|EPI_ISL_510660\|2 | ···················· | ···················· | ···················· | ···················· | ···················· | ···················· |
| F | 1244 hCoV-19/South Korea/KCDC2325/2020\|EPI_ISL_510661\|2 | ···················· | ···················· | ···················· | ···················· | ···················· | ···················· |
| F | 1245 hCoV-19/South Korea/KCDC2326/2020\|EPI_ISL_510662\|2 | ···················· | ···················· | ···················· | ···················· | ···················· | ···················· |
| F | 1246 hCoV-19/South Korea/KCDC2327/2020\|EPI_ISL_510663\|2 | ···················· | ···················· | ···················· | ···················· | ···················· | ···················· |
| F | 1247 hCoV-19/South Korea/KCDC2328/2020\|EPI_ISL_510664\|2 | ···················· | ···················· | ···················· | ···················· | ···················· | ···················· |
| F | 1248 hCoV-19/South Korea/KCDC2329/2020\|EPI_ISL_510665\|2 | ···················· | ···················· | ···················· | ···················· | ···················· | ···················· |
| F | 1249 hCoV-19/South Korea/KCDC2336/2020\|EPI_ISL_510672\|2 | ···················· | ···················· | ···················· | ···················· | ···················· | ···················· |
| F | 1250 hCoV-19/England/BRIS-13259C/2020\|EPI_ISL_511976\|20 | ···················· | ···················· | ···················· | ···················· | ···················· | ···················· |
| F | 1251 hCoV-19/England/BRIS-132E19/2020\|EPI_ISL_511978\|20 | ···················· | ···················· | ···················· | ···················· | ···················· | ···················· |
| F | 1252 hCoV-19/England/BRIS-132CB5/2020\|EPI_ISL_511979\|20 | ···················· | ···················· | ···················· | ···················· | ···················· | ···················· |
| F | 1253 hCoV-19/England/BRIS-132F61/2020\|EPI_ISL_511980\|20 | ···················· | ·············A······ | ···················· | ···················· | ···················· | ···················· |
| F | 1254 hCoV-19/England/BRIS-132D67/2020\|EPI_ISL_511981\|20 | ···················· | ···················· | ···················· | ···················· | ·········T·········· | ···················· |
| F | 1255 hCoV-19/Switzerland/TG-ETHZ-210010/2020\|EPI_ISL_51 | ···················· | ···················· | ···················· | ···················· | ···················· | ···················· |
| F | 1256 hCoV-19/Switzerland/AG-ETHZ-210017/2020\|EPI_ISL_51 | ···················· | ···················· | ···················· | ···················· | ···················· | ···················· |
| F | 1257 hCoV-19/Switzerland/ZH-ETHZ-210024/2020\|EPI_ISL_51 | ···················· | ···················· | ···················· | ···················· | ···················· | ···················· |
| F | 1258 hCoV-19/Switzerland/ZH-ETHZ-210031/2020\|EPI_ISL_51 | ···················· | ···················· | ···················· | ···················· | ···················· | ···················· |
| F | 1259 hCoV-19/Switzerland/ZH-ETHZ-210044/2020\|EPI_ISL_51 | ···················· | ···················· | ···················· | ···················· | ···················· | ···················· |
| F | 1260 hCoV-19/Switzerland/VD-ETHZ-210053/2020\|EPI_ISL_51 | ···················· | ···················· | ···················· | ···················· | ···················· | ···················· |
| F | 1261 hCoV-19/Switzerland/ZH-ETHZ-210057/2020\|EPI_ISL_51 | ···················· | ···················· | ···················· | ···················· | ···················· | ···················· |
| F | 1262 hCoV-19/Switzerland/BE-ETHZ-210076/2020\|EPI_ISL_51 | ···················· | ···················· | ···················· | ···················· | ···················· | ···················· |
| F | 1263 hCoV-19/Switzerland/ZH-ETHZ-210080/2020\|EPI_ISL_51 | ···················· | ···················· | ···················· | ···················· | ···················· | ···················· |
| F | 1264 hCoV-19/Switzerland/BL-ETHZ-220001/2020\|EPI_ISL_51 | ···················· | ···················· | ···················· | ···················· | ···················· | ···················· |
| F | 1265 hCoV-19/USA/FL-BPHL-0855/2020\|EPI_ISL_514201\|2020- | ···················· | ···················· | ···················· | ···················· | ···················· | ···················· |
| F | 1266 hCoV-19/USA/FL-BPHL-0856/2020\|EPI_ISL_514202\|2020- | ···················· | ···················· | ···················· | ···················· | ···················· | ···················· |
| F | 1267 hCoV-19/USA/FL-BPHL-0857/2020\|EPI_ISL_514203\|2020- | ···················· | ···················· | ···················· | ···················· | ···················· | ···················· |
| F | 1268 hCoV-19/USA/FL-BPHL-0858/2020\|EPI_ISL_514204\|2020- | ···················· | ···················· | ···················· | ···················· | ···················· | ···················· |
| F | 1269 hCoV-19/USA/FL-BPHL-0859/2020\|EPI_ISL_514205\|2020- | ···················· | ···················· | ···················· | ···················· | ···················· | ···················· |
| F | 1270 hCoV-19/USA/FL-BPHL-0861/2020\|EPI_ISL_514207\|2020- | ···················· | ···················· | ···················· | ···················· | ···················· | ···················· |

| | | | | | | | | |
|---|---|---|---|---|---|---|---|---|
| F | 1271 | hCoV-19/Brazil/SP-118/2020\|EPI_ISL_515520\|2020-03- | ···················· | ···················· | ···················· | ···················· | ···················· | ···················· |
| F | 1272 | hCoV-19/Brazil/SP-120/2020\|EPI_ISL_515521\|2020-03- | ···················· | ···················· | ···················· | ···················· | ···················· | ···················· |
| F | 1273 | hCoV-19/Brazil/SP-122/2020\|EPI_ISL_515522\|2020-03- | ···················· | ···················· | ···················· | ···················· | ···················· | ···················· |
| F | 1274 | hCoV-19/Brazil/SP-124/2020\|EPI_ISL_515523\|2020-04- | ···················· | ···················· | ···················· | ···················· | ···················· | ···················· |
| F | 1275 | hCoV-19/Brazil/SP-125/2020\|EPI_ISL_515524\|2020-04- | ···················· | ···················· | ···················· | ···················· | ···················· | ···················· |
| F | 1276 | hCoV-19/Brazil/SP-600/2020\|EPI_ISL_515525\|2020-06- | ···················· | ···················· | ···················· | ···················· | ···················· | ···················· |
| F | 1277 | hCoV-19/Brazil/SP-602/2020\|EPI_ISL_515526\|2020-03- | ···················· | ···················· | ···················· | ···················· | ···················· | ···················· |
| F | 1278 | hCoV-19/Brazil/SP-624/2020\|EPI_ISL_515527\|2020-03- | ···················· | ···················· | ···················· | ···················· | ···················· | ···················· |
| F | 1279 | hCoV-19/Brazil/SP-625/2020\|EPI_ISL_515528\|2020-03- | ···················· | ···················· | ···················· | ···················· | ···················· | ···················· |
| F | 1280 | hCoV-19/Brazil/SP-626/2020\|EPI_ISL_515529\|2020-03- | ···················· | ···················· | ···················· | ···················· | ···················· | ···················· |
| F | 1281 | hCoV-19/USA/WA-S2668/2020\|EPI_ISL_497300\|2020-06-2 | ···················· | ···················· | ···················· | ···················· | ···················· | ···················· |
| F | 1282 | hCoV-19/USA/WA-S2669/2020\|EPI_ISL_497301\|2020-06-2 | ···················· | ···················· | ···················· | ···················· | ···················· | ···················· |
| F | 1283 | hCoV-19/USA/WA-S2670/2020\|EPI_ISL_497302\|2020-06-2 | ···················· | ···················· | ···················· | ···················· | ···················· | ···················· |
| F | 1284 | hCoV-19/USA/WA-S2671/2020\|EPI_ISL_497303\|2020-06-2 | ···················· | ···················· | ···················· | ···················· | ···················· | ···················· |
| F | 1285 | hCoV-19/USA/WA-S2672/2020\|EPI_ISL_497304\|2020-06-2 | ···················· | ···················· | ···················· | ···················· | ···················· | ···················· |
| F | 1286 | hCoV-19/USA/WA-S2673/2020\|EPI_ISL_497305\|2020-06-2 | ···················· | ···················· | ···················· | ···················· | ···················· | ···················· |
| F | 1287 | hCoV-19/USA/WA-S2674/2020\|EPI_ISL_497306\|2020-06-2 | ···················· | ···················· | ···················· | ···················· | ···················· | ···················· |
| F | 1288 | hCoV-19/USA/WA-S2675/2020\|EPI_ISL_497307\|2020-06-2 | ···················· | ···················· | ···················· | ···················· | ···················· | ···················· |
| F | 1289 | hCoV-19/USA/WA-S2676/2020\|EPI_ISL_497308\|2020-06-2 | ···················· | ···················· | ···················· | ···················· | ···················· | ···················· |
| F | 1290 | hCoV-19/USA/WA-S2678/2020\|EPI_ISL_497310\|2020-06-2 | ···················· | ···················· | ···················· | ···················· | ···················· | ···················· |
| F | 1291 | hCoV-19/USA/WA-S2679/2020\|EPI_ISL_497311\|2020-06-2 | ···················· | ···················· | ···················· | ···················· | ···················· | ···················· |
| F | 1292 | hCoV-19/USA/WA-S2680/2020\|EPI_ISL_497312\|2020-06-2 | ···················· | ···················· | ···················· | ···················· | ···················· | ···················· |
| F | 1293 | hCoV-19/USA/WA-S2681/2020\|EPI_ISL_497313\|2020-06-2 | ···················· | ···················· | ···················· | ···················· | ···················· | ···················· |
| F | 1294 | hCoV-19/USA/WA-S2683/2020\|EPI_ISL_497315\|2020-06-2 | ···················· | ···················· | ···················· | ···················· | ···················· | ···················· |
| F | 1295 | hCoV-19/USA/WA-S2684/2020\|EPI_ISL_497316\|2020-06-2 | ···················· | ···················· | ···················· | ···················· | ···················· | ···················· |
| F | 1296 | hCoV-19/USA/WA-S2685/2020\|EPI_ISL_497317\|2020-06-2 | ···················· | ···················· | ···················· | ···················· | ···················· | ···················· |
| F | 1297 | hCoV-19/USA/WA-S2686/2020\|EPI_ISL_497318\|2020-06-2 | ···················· | ···················· | ···················· | ···················· | ···················· | ···················· |
| F | 1298 | hCoV-19/USA/WA-S2687/2020\|EPI_ISL_497319\|2020-06-2 | ···················· | ···················· | ···················· | ···················· | ···················· | ···················· |
| F | 1299 | hCoV-19/USA/WA-S2689/2020\|EPI_ISL_497321\|2020-06-2 | ···················· | ···················· | ···················· | ···················· | ···················· | ···················· |
| F | 1300 | hCoV-19/USA/WA-S2690/2020\|EPI_ISL_497322\|2020-06-2 | ···················· | ···················· | ···················· | ···················· | ···················· | ···················· |
| F | 1301 | hCoV-19/Singapore/703/2020\|EPI_ISL_498601\|2020-06- | ···················· | ···················· | ···················· | ···················· | ···················· | ···················· |
| F | 1302 | hCoV-19/Singapore/704/2020\|EPI_ISL_498602\|2020-06- | ···················· | ···················· | ···················· | ···················· | ···················· | ···················· |
| F | 1303 | hCoV-19/Singapore/705/2020\|EPI_ISL_498603\|2020-06- | ···················· | ···················· | ···················· | ···················· | ···················· | ···················· |
| F | 1304 | hCoV-19/Singapore/706/2020\|EPI_ISL_498604\|2020-06- | ···················· | ···················· | ···················· | ···················· | ···················· | ···················· |
| F | 1305 | hCoV-19/Singapore/707/2020\|EPI_ISL_498605\|2020-06- | ···················· | ···················· | ···················· | ···················· | ···················· | ···················· |
| F | 1306 | hCoV-19/Singapore/710/2020\|EPI_ISL_498608\|2020-07- | ···················· | ···················· | ···················· | ···················· | ···················· | ···················· |
| F | 1307 | hCoV-19/Singapore/711/2020\|EPI_ISL_498609\|2020-07- | ···················· | ···················· | ···················· | ···················· | ···················· | ···················· |
| F | 1308 | hCoV-19/Singapore/712/2020\|EPI_ISL_498610\|2020-07- | ···················· | ···················· | ···················· | ···················· | ···················· | ···················· |
| F | 1309 | hCoV-19/Singapore/713/2020\|EPI_ISL_498611\|2020-07- | ···················· | ···················· | ···················· | ···················· | ···················· | ···················· |
| F | 1310 | hCoV-19/Singapore/715/2020\|EPI_ISL_498613\|2020-07- | ···················· | ···················· | ···················· | ···················· | ···················· | ···················· |
| F | 1311 | hCoV-19/Singapore/717/2020\|EPI_ISL_498615\|2020-07- | ···················· | ···················· | ···················· | ···················· | ···················· | ···················· |
| F | 1312 | hCoV-19/Singapore/718/2020\|EPI_ISL_498616\|2020-07- | ···················· | ···················· | ···················· | ···················· | ···················· | ···················· |
| F | 1313 | hCoV-19/Singapore/720/2020\|EPI_ISL_498618\|2020-07- | ···················· | ···················· | ···················· | ···················· | ···················· | ···················· |
| F | 1314 | hCoV-19/Switzerland/SZ-ETHZ-190002/2020\|EPI_ISL_49 | ···················· | ···················· | ···················· | ···················· | ···················· | ···················· |
| F | 1315 | hCoV-19/Switzerland/SZ-ETHZ-190003/2020\|EPI_ISL_49 | ···················· | ···················· | ···················· | ···················· | ···················· | ···················· |
| F | 1316 | hCoV-19/Switzerland/ZH-ETHZ-190004/2020\|EPI_ISL_49 | ···················· | ···················· | ···················· | ···················· | ···················· | ···················· |
| F | 1317 | hCoV-19/Switzerland/ZH-ETHZ-190008/2020\|EPI_ISL_49 | ···················· | ···················· | ···················· | ···················· | ···················· | ···················· |
| F | 1318 | hCoV-19/Switzerland/BE-ETHZ-190010/2020\|EPI_ISL_49 | ···················· | ···················· | ···················· | ···················· | ···················· | ···················· |
| F | 1319 | hCoV-19/Belgium/ULG-10246/2020\|EPI_ISL_498628\|2020 | ···················· | ···················· | ···················· | ···················· | ···················· | ···················· |
| F | 1320 | hCoV-19/Belgium/ULG-10247/2020\|EPI_ISL_498629\|2020 | ···················· | ···················· | ···················· | ···················· | ···················· | ···················· |
| F | 1321 | hCoV-19/USA/UT-01856/2020\|EPI_ISL_498630\|2020-04-1 | ···················· | ···················· | ···················· | ···················· | ···················· | ···················· |
| F | 1322 | hCoV-19/USA/UT-01857/2020\|EPI_ISL_498631\|2020-04-1 | ···················· | ···················· | ···················· | ···················· | ···················· | ···················· |
| F | 1323 | hCoV-19/USA/UT-01859/2020\|EPI_ISL_498632\|2020-04-1 | ···················· | ···················· | ···················· | ···················· | ···················· | ···················· |
| F | 1324 | hCoV-19/USA/UT-01860/2020\|EPI_ISL_498633\|2020-04-1 | ···················· | ···················· | ···················· | ···················· | ···················· | ···················· |
| F | 1325 | hCoV-19/USA/UT-01861/2020\|EPI_ISL_498634\|2020-04-1 | ···················· | ···················· | ···················· | ···················· | ···················· | ···················· |
| F | 1326 | hCoV-19/USA/UT-01862/2020\|EPI_ISL_498635\|2020-04-1 | ···················· | ···················· | ···················· | ···················· | ···················· | ···················· |
| F | 1327 | hCoV-19/USA/UT-01864/2020\|EPI_ISL_498637\|2020-04-1 | ···················· | ···················· | ···················· | ···················· | ···················· | ···················· |
| F | 1328 | hCoV-19/USA/UT-01865/2020\|EPI_ISL_498638\|2020-04-1 | ···················· | ···················· | ···················· | ···················· | ···················· | ···················· |
| F | 1329 | hCoV-19/USA/UT-01866/2020\|EPI_ISL_498639\|2020-04-1 | ···················· | ···················· | ···················· | ···················· | ···················· | ···················· |
| F | 1330 | hCoV-19/USA/UT-03317/2020\|EPI_ISL_498640\|2020-04-2 | ···················· | ···················· | ···················· | ···················· | ···················· | ···················· |
| F | 1331 | hCoV-19/USA/UT-03320/2020\|EPI_ISL_498641\|2020-04-2 | ···················· | ···················· | ···················· | ···················· | ···················· | ···················· |
| F | 1332 | hCoV-19/USA/UT-03325/2020\|EPI_ISL_498642\|2020-04-2 | ···················· | ···················· | ···················· | ···················· | ···················· | ···················· |
| F | 1333 | hCoV-19/USA/UT-03328/2020\|EPI_ISL_498643\|2020-04-2 | ···················· | ···················· | ···················· | ···················· | ···················· | ···················· |
| F | 1334 | hCoV-19/USA/UT-03332/2020\|EPI_ISL_498644\|2020-04-2 | ···················· | ···················· | ···················· | ···················· | ···················· | ···················· |
| F | 1335 | hCoV-19/USA/UT-03338/2020\|EPI_ISL_498645\|2020-04-2 | ···················· | ···················· | ···················· | ···················· | ···················· | ···················· |
| F | 1336 | hCoV-19/USA/UT-03341/2020\|EPI_ISL_498646\|2020-04-2 | ···················· | ···················· | ···················· | ···················· | ···················· | ···················· |
| F | 1337 | hCoV-19/USA/UT-03345/2020\|EPI_ISL_498647\|2020-04-2 | ···················· | ···················· | ···················· | ···················· | ···················· | ···················· |
| F | 1338 | hCoV-19/USA/UT-03348/2020\|EPI_ISL_498648\|2020-04-2 | ···················· | ···················· | ···················· | ···················· | ···················· | ···················· |
| F | 1339 | hCoV-19/USA/UT-03354/2020\|EPI_ISL_498649\|2020-04-2 | ···················· | ···················· | ···················· | ···················· | ···················· | ···················· |
| F | 1340 | hCoV-19/USA/UT-03356/2020\|EPI_ISL_498650\|2020-04-2 | ···················· | ···················· | ···················· | ···················· | ···················· | ···················· |
| F | 1341 | hCoV-19/USA/UT-03357/2020\|EPI_ISL_498651\|2020-04-2 | ···················· | ···················· | ···················· | ···················· | ···················· | ···················· |
| F | 1342 | hCoV-19/USA/UT-03366/2020\|EPI_ISL_498652\|2020-04-2 | ···················· | ···················· | ···················· | ···················· | ···················· | ···················· |
| F | 1343 | hCoV-19/USA/UT-03368/2020\|EPI_ISL_498653\|2020-04-2 | ···················· | ···················· | ···················· | ···················· | ···················· | ···················· |
| F | 1344 | hCoV-19/England/CAMB-1B5467/2020\|EPI_ISL_499968\|20 | ···················· | ···················· | ···················· | ···················· | ···················· | ···················· |
| F | 1345 | hCoV-19/England/CAMB-1B55FB/2020\|EPI_ISL_499969\|20 | ···················· | ···················· | ···················· | ···················· | ···················· | ···················· |
| F | 1346 | hCoV-19/England/CAMB-1B5607/2020\|EPI_ISL_499970\|20 | ···················· | ···················· | ···················· | ···················· | ···················· | ···················· |
| F | 1347 | hCoV-19/England/CAMB-1B5616/2020\|EPI_ISL_499971\|20 | ···················· | ···················· | ···················· | ···················· | ···················· | ···················· |
| F | 1348 | hCoV-19/England/CAMB-1B5643/2020\|EPI_ISL_499972\|20 | ···················· | ···················· | ···················· | ···················· | ···················· | ···················· |
| F | 1349 | hCoV-19/England/CAMB-1B5670/2020\|EPI_ISL_499973\|20 | ···················· | ···················· | ···················· | ···················· | ···················· | ···················· |
| F | 1350 | hCoV-19/England/LCST-24D71C9/2020\|EPI_ISL_499974\|2 | ···················· | ···················· | ···················· | ···················· | ···················· | ···················· |

F 1351 hCoV-19/England/LCST-24D71D8/2020|EPI_ISL_499975|2 ···················· ···················· ····················· ···················· ···················· ····················
F 1352 hCoV-19/England/LCST-24D725D/2020|EPI_ISL_499976|2 ···················· ···················· ····················· ···················· ···················· ····················
F 1353 hCoV-19/England/LCST-24D727B/2020|EPI_ISL_499977|2 ···················· ···················· ····················· ···················· ···················· ····················
F 1354 hCoV-19/England/LCST-24D72B7/2020|EPI_ISL_499978|2 ···················· ···················· ····················· ···················· ···················· ····················
F 1355 hCoV-19/England/LCST-24D732D/2020|EPI_ISL_499979|2 ···················· ···················· ····················· ···················· ···················· ····················
F 1356 hCoV-19/England/LCST-24D733C/2020|EPI_ISL_499980|2 ···················· ···················· ····················· ···················· ···················· ····················
F 1357 hCoV-19/England/LCST-24D734B/2020|EPI_ISL_499981|2 ···················· ···················· ····················· ···················· ···················· ····················
F 1358 hCoV-19/England/LCST-24D7387/2020|EPI_ISL_499982|2 ···················· ···················· ····················· ···················· ···················· ····················
F 1359 hCoV-19/England/LCST-24D741B/2020|EPI_ISL_499983|2 ···················· ···················· ····················· ···················· ···················· ····················
F 1361 hCoV-19/England/LOND-124D2DF/2020|EPI_ISL_507080|2 ···················· ···················· ····················· ···················· ···················· ····················
F 1362 hCoV-19/England/LOND-124D2EE/2020|EPI_ISL_507081|2 ···················· ···················· ····················· ···················· ···················· ····················
F 1363 hCoV-19/England/LOND-124D2FD/2020|EPI_ISL_507082|2 ···················· ···················· ····················· ···················· ···················· ····················
F 1364 hCoV-19/England/LOND-124D309/2020|EPI_ISL_507083|2 ···················· ···················· ····················· ···················· ···················· ····················
F 1365 hCoV-19/England/LOND-124D318/2020|EPI_ISL_507084|2 ···················· ···················· ····················· ···················· ···················· ····················
F 1366 hCoV-19/England/LOND-124D345/2020|EPI_ISL_507086|2 ···················· ···················· ····················· ···················· ···················· ····················
F 1367 hCoV-19/England/LOND-124D354/2020|EPI_ISL_507087|2 ···················· ···················· ····················· ···················· ···················· ····················
F 1368 hCoV-19/England/LOND-124D372/2020|EPI_ISL_507089|2 ···················· ···················· ····················· ···················· ···················· ····················
F 1369 hCoV-19/England/LOND-124D381/2020|EPI_ISL_507090|2 ···················· ···················· ····················· ···················· ···················· ····················
F 1371 hCoV-19/England/LOND-124D3AF/2020|EPI_ISL_507092|2 ···················· ···················· ····················· ···················· ···················· ····················
F 1372 hCoV-19/England/LOND-124D3BE/2020|EPI_ISL_507093|2 ···················· ···················· ····················· ···················· ···················· ····················
F 1374 hCoV-19/England/LOND-124D3FA/2020|EPI_ISL_507096|2 ···················· ···················· ····················· ···················· ···················· ····················
F 1376 hCoV-19/England/LOND-124D451/2020|EPI_ISL_507098|2 ···················· ···················· ····················· ···················· ···················· ····················
F 1377 hCoV-19/England/LOND-124D4CA/2020|EPI_ISL_507103|2 ···················· ···················· ····················· ···················· ···················· ····················
F 1378 hCoV-19/England/LOND-124D4E8/2020|EPI_ISL_507104|2 ···················· ···················· ····················· ···················· ···················· ····················
F 1379 hCoV-19/England/LOND-124D503/2020|EPI_ISL_507105|2 ···················· ···················· ····················· ···················· ···················· ····················
F 1380 hCoV-19/England/LOND-124D54F/2020|EPI_ISL_507107|2 ···················· ···················· ····················· ···················· ···················· ····················
F 1381 hCoV-19/England/LOND-124D56D/2020|EPI_ISL_507108|2 ···················· ···················· ····················· ···················· ···················· ····················
F 1382 hCoV-19/England/NORT-299F3A/2020|EPI_ISL_507110|20 ···················· ···················· ····················· ···················· ···················· ····················
F 1383 hCoV-19/England/NORT-29A0CD/2020|EPI_ISL_507111|20 ···················· ···················· ····················· ···················· ···················· ····················
F 1384 hCoV-19/England/NORT-29A1D9/2020|EPI_ISL_507112|20 ···················· ···················· ····················· ···················· ···················· ····················
F 1385 hCoV-19/England/NORT-29A388/2020|EPI_ISL_507113|20 ···················· ···················· ····················· ···················· ···················· ····················
F 1386 hCoV-19/England/NORT-29A731/2020|EPI_ISL_507116|20 ···················· ···················· ····················· ···················· ···················· ····················
F 1387 hCoV-19/England/NORT-29A740/2020|EPI_ISL_507117|20 ···················· ···················· ····················· ···················· ···················· ····················
F 1388 hCoV-19/England/NORT-29A79B/2020|EPI_ISL_507118|20 ···················· ···················· ····················· ···················· ···················· ····················
F 1389 hCoV-19/India/MH-MW26/2020|EPI_ISL_508430|2020-05- ···················· ···················· ····················· ···················· ···················· ····················
F 1390 hCoV-19/India/MH-MW28/2020|EPI_ISL_508431|2020-05- ···················· ···················· ····················· ···················· ···················· ····················
F 1391 hCoV-19/India/MH-MW29/2020|EPI_ISL_508432|2020-05- ···················· ···················· ····················· ···················· ···················· ····················
F 1392 hCoV-19/India/MH-MW3/2020|EPI_ISL_508433|2020-04-2 ···················· ···················· ····················· ···················· ···················· ····················
F 1393 hCoV-19/India/OR-MW30/2020|EPI_ISL_508434|2020-05- ···················· ···················· ····················· ···················· ···················· ····················
F 1394 hCoV-19/India/MH-MW32/2020|EPI_ISL_508435|2020-05- ···················· ···················· ····················· ···················· ···················· ····················
F 1395 hCoV-19/India/MH-MW33/2020|EPI_ISL_508436|2020-05- ···················· ···················· ····················· ···················· ···················· ····················
F 1396 hCoV-19/India/MH-MW34/2020|EPI_ISL_508437|2020-06- ···················· ·A·················· ····················· ···················· ···················· ··A·················
F 1397 hCoV-19/India/MH-MW38/2020|EPI_ISL_508438|2020-06- ···················· ···················· ····················· ···················· ···················· ····················
F 1398 hCoV-19/India/MH-MW5/2020|EPI_ISL_508439|2020-04-2 ···················· ···················· ····················· ···················· ···················· ····················
F 1399 hCoV-19/India/MH-MW8/2020|EPI_ISL_508440|2020-04-2 ···················· ···················· ····················· ···················· ···················· ····················
F 1400 hCoV-19/India/WB-S35/2020|EPI_ISL_508444|2020-05-0 ···················· ···················· ····················· ···················· ···················· ····················
F 1401 hCoV-19/India/WB-S38/2020|EPI_ISL_508446|2020-05-0 ···················· ···················· ····················· ···················· ···················· ····················
F 1402 hCoV-19/India/WB-S39/2020|EPI_ISL_508447|2020-04-2 ···················· ···················· ····················· ···················· ···················· ····················
F 1403 hCoV-19/India/WB-S40/2020|EPI_ISL_508448|2020-05-0 ···················· ···················· ····················· ···················· ···················· ····················
F 1404 hCoV-19/India/WB-S41/2020|EPI_ISL_508449|2020-05-0 ···················· ···················· ····················· ···················· ···················· ····················
F 1405 hCoV-19/USA/FL-BPHL-0702/2020|EPI_ISL_509760|2020- ···················· ···················· ····················· ···················· ···················· ····················
F 1406 hCoV-19/USA/FL-BPHL-0703/2020|EPI_ISL_509761|2020- ···················· ···················· ····················· ···················· ···················· ····················
F 1407 hCoV-19/USA/FL-BPHL-0704/2020|EPI_ISL_509762|2020- ···················· ···················· ····················· ···················· ···················· ····················
F 1408 hCoV-19/USA/FL-BPHL-0705/2020|EPI_ISL_509763|2020- ···················· ···················· ····················· ···················· ···················· ····················
F 1409 hCoV-19/USA/FL-BPHL-0706/2020|EPI_ISL_509764|2020- ···················· ···················· ····················· ···················· ···················· ····················
F 1410 hCoV-19/USA/FL-BPHL-0707/2020|EPI_ISL_509765|2020- ···················· ···················· ····················· ···················· ···················· ····················
F 1411 hCoV-19/USA/FL-BPHL-0708/2020|EPI_ISL_509766|2020- ···················· ···················· ····················· ···················· ···················· ····················
F 1412 hCoV-19/USA/FL-BPHL-0709/2020|EPI_ISL_509767|2020- ···················· ···················· ····················· ···················· ···················· ····················
F 1413 hCoV-19/USA/FL-BPHL-0711/2020|EPI_ISL_509769|2020- ···················· ···················· ····················· ···················· ···················· ····················
F 1414 hCoV-19/USA/FL-BPHL-0713/2020|EPI_ISL_509771|2020- ···················· ···················· ····················· ···················· ···················· ····················
F 1415 hCoV-19/USA/FL-BPHL-0714/2020|EPI_ISL_509772|2020- ···················· ···················· ····················· ···················· ···················· ····················
F 1416 hCoV-19/USA/FL-BPHL-0715/2020|EPI_ISL_509773|2020- ···················· ···················· ····················· ···················· ···················· ····················
F 1417 hCoV-19/USA/FL-BPHL-0716/2020|EPI_ISL_509774|2020- ···················· ···················· ····················· ···················· ···················· ····················
F 1418 hCoV-19/USA/FL-BPHL-0718/2020|EPI_ISL_509776|2020- ···················· ···················· ····················· ···················· ···················· ····················
F 1419 hCoV-19/USA/FL-BPHL-0720/2020|EPI_ISL_509778|2020- ···················· ···················· ····················· ···················· ···················· ····················
F 1420 hCoV-19/Switzerland/BE-ETHZ-200061/2020|EPI_ISL_51 ···········T········ ···················· ····················· ···················· ···················· ····················
F 1421 hCoV-19/Switzerland/BL-ETHZ-200062/2020|EPI_ISL_51 ···················· ···················· ····················· ···················· ···················· ····················
F 1422 hCoV-19/Switzerland/ZH-ETHZ-200064/2020|EPI_ISL_51 ···················· ···················· ····················· ···················· ···················· ····················
F 1423 hCoV-19/Switzerland/VD-ETHZ-200065/2020|EPI_ISL_51 ···················· ···················· ····················· ···················· ···················· ····················
F 1424 hCoV-19/Switzerland/BS-ETHZ-200066/2020|EPI_ISL_51 ···················· ·R·················· ····················· ···················· ···················· ····················
F 1425 hCoV-19/Switzerland/GE-ETHZ-200067/2020|EPI_ISL_51 ···················· ···················· ····················· ···················· ···················· ····················
F 1426 hCoV-19/Switzerland/SG-ETHZ-200068/2020|EPI_ISL_51 ···················· ···················· ····················· ···················· ···················· ····················
F 1427 hCoV-19/Switzerland/BL-ETHZ-210006/2020|EPI_ISL_51 ···················· ···················· ····················· ···················· ···················· ····················
F 1428 hCoV-19/Switzerland/BL-ETHZ-210007/2020|EPI_ISL_51 ···················· ···················· ····················· ···················· ···················· ····················
F 1429 hCoV-19/Switzerland/BL-ETHZ-210008/2020|EPI_ISL_51 ···················· ···················· ····················· ···················· ···················· ····················
F 1430 hCoV-19/Switzerland/LU-ETHZ-210009/2020|EPI_ISL_51 ···················· ···················· ····················· ···················· ···················· ····················
F 1431 hCoV-19/Switzerland/ZH-ETHZ-210011/2020|EPI_ISL_51 ···················· ···················· ····················· ···················· ···················· ····················
F 1432 hCoV-19/Switzerland/ZH-ETHZ-210012/2020|EPI_ISL_51 ···················· ···················· ····················· ···················· ···················· ····················
F 1433 hCoV-19/Switzerland/ZH-ETHZ-210013/2020|EPI_ISL_51 ···················· ···················· ····················· ···················· ···················· ····················
F 1434 hCoV-19/Switzerland/AG-ETHZ-210014/2020|EPI_ISL_51 ···················· ···················· ····················· ···················· ···················· ····················

| | | | | | | | |
|---|---|---|---|---|---|---|---|
| F | 1435 hCoV-19/Switzerland/AG-ETHZ-210015/2020\|EPI_ISL_51 | ···················· | ···················· | ···················· | ···················· | ···················· | ···················· |
| F | 1436 hCoV-19/Switzerland/BL-ETHZ-210018/2020\|EPI_ISL_51 | ···················· | ···················· | ···················· | ···················· | ···················· | ···················· |
| F | 1437 hCoV-19/USA/FL-BPHL-0865/2020\|EPI_ISL_514211\|2020- | ···················· | ···················· | ···················· | ···················· | ···················· | ···················· |
| F | 1438 hCoV-19/USA/FL-BPHL-0866/2020\|EPI_ISL_514212\|2020- | ···················· | ···················· | ···················· | ···················· | ···················· | ···················· |
| F | 1439 hCoV-19/USA/FL-BPHL-0867/2020\|EPI_ISL_514213\|2020- | ···················· | ···················· | ···················· | ···················· | ···················· | ···················· |
| F | 1440 hCoV-19/USA/FL-BPHL-0868/2020\|EPI_ISL_514214\|2020- | ···················· | ···················· | ···················· | ···················· | ···················· | ···················· |
| F | 1441 hCoV-19/USA/FL-BPHL-0869/2020\|EPI_ISL_514215\|2020- | ···················· | ···················· | ···················· | ···················· | ···················· | ···················· |
| F | 1442 hCoV-19/USA/FL-BPHL-0870/2020\|EPI_ISL_514216\|2020- | ···················· | ···················· | ···················· | ···················· | ···················· | ···················· |
| F | 1443 hCoV-19/USA/FL-BPHL-0873/2020\|EPI_ISL_514219\|2020- | ···················· | ···················· | ···················· | ···················· | ···················· | ···················· |
| F | 1444 hCoV-19/USA/FL-BPHL-0874/2020\|EPI_ISL_514220\|2020- | ···················· | ···················· | ···················· | ···················· | ···················· | ···················· |
| F | 1445 hCoV-19/USA/FL-BPHL-0876/2020\|EPI_ISL_514222\|2020- | ···················· | ···················· | ···················· | ···················· | ···················· | ···················· |
| F | 1446 hCoV-19/USA/FL-BPHL-0877/2020\|EPI_ISL_514223\|2020- | ···················· | ···A················ | ···················· | ···················· | ···················· | ···················· |
| F | 1447 hCoV-19/USA/FL-BPHL-0878/2020\|EPI_ISL_514224\|2020- | ···················· | ···A················ | ···················· | ···················· | ···················· | ···················· |
| F | 1448 hCoV-19/USA/FL-BPHL-0879/2020\|EPI_ISL_514225\|2020- | ···················· | ···················· | ···················· | ···················· | ···················· | ···················· |
| F | 1449 hCoV-19/Peru/LIM-INS-041/2020\|EPI_ISL_514226\|2020- | ···················· | ···················· | ···················· | ···················· | ···················· | ···················· |
| F | 1450 hCoV-19/Peru/LIM-INS-042/2020\|EPI_ISL_514227\|2020- | ···················· | ···················· | ···················· | ···················· | ···················· | ···················· |
| F | 1451 hCoV-19/Bangladesh/BCSIR-NILMRC-350/2020\|EPI_ISL_5 | ···················· | ···················· | ···················· | ···················· | ···················· | ···················· |
| F | 1452 hCoV-19/Bangladesh/BCSIR-NILMRC-351/2020\|EPI_ISL_5 | ···················· | ·A·················· | ···················· | ···················· | ···················· | ··A················· |
| F | 1453 hCoV-19/Bangladesh/BCSIR-NILMRC-353/2020\|EPI_ISL_5 | ···················· | ···················· | ···················· | ···················· | ···················· | ···················· |
| F | 1454 hCoV-19/Bangladesh/BCSIR-NILMRC-354/2020\|EPI_ISL_5 | ···················· | ···················· | ···················· | ···················· | ···················· | ···················· |
| F | 1455 hCoV-19/Bangladesh/BCSIR-NILMRC-359/2020\|EPI_ISL_5 | ···················· | ···················· | ···················· | ···················· | ···················· | ···················· |
| F | 1456 hCoV-19/Bangladesh/BCSIR-NILMRC-361/2020\|EPI_ISL_5 | ···················· | ···················· | ···················· | ···················· | ···················· | ···················· |
| F | 1457 hCoV-19/Bangladesh/BCSIR-NILMRC-362/2020\|EPI_ISL_5 | ···················· | ···················· | ···················· | ···················· | ···················· | ···················· |
| F | 1458 hCoV-19/Bangladesh/BCSIR-NILMRC-363/2020\|EPI_ISL_5 | ···················· | ···················· | ···················· | ···················· | ···················· | ···················· |
| F | 1459 hCoV-19/Bangladesh/BCSIR-NILMRC-364/2020\|EPI_ISL_5 | ·······T············ | ···················· | ···················· | ···················· | ···················· | ···················· |
| F | 1460 hCoV-19/Bangladesh/BCSIR-NILMRC-365/2020\|EPI_ISL_5 | ···················· | ···················· | ···················· | ···················· | ···················· | ···················· |
| F | 1461 hCoV-19/Bangladesh/BCSIR-NILMRC-367/2020\|EPI_ISL_5 | ···················· | ···················· | ···················· | ···················· | ···················· | ···················· |
| F | 1462 hCoV-19/Bangladesh/BCSIR-NILMRC-368/2020\|EPI_ISL_5 | ···················· | ···················· | ···················· | ···················· | ···················· | ···················· |
| F | 1463 hCoV-19/Bangladesh/BCSIR-NILMRC-370/2020\|EPI_ISL_5 | ···················· | ···················· | ···················· | ···················· | ···················· | ···················· |
| F | 1464 hCoV-19/Bangladesh/BCSIR-NILMRC-385/2020\|EPI_ISL_5 | ···················· | ···················· | ···················· | ···················· | ···················· | ···················· |
| F | 1465 hCoV-19/Bangladesh/BCSIR-NILMRC-386/2020\|EPI_ISL_5 | ···················· | ···················· | ···················· | ···················· | ···················· | ···················· |
| F | 1466 hCoV-19/Bangladesh/BCSIR-NILMRC-387/2020\|EPI_ISL_5 | ···················· | ···················· | ···················· | ···················· | ···················· | ···················· |
| F | 1467 hCoV-19/Bangladesh/BCSIR-NILMRC-369/2020\|EPI_ISL_5 | ···················· | ···················· | ···················· | ···················· | ···················· | ···················· |
| F | 1468 hCoV-19/Bangladesh/BCSIR-NILMRC-381/2020\|EPI_ISL_5 | ···················· | ·A·················· | ···················· | ···················· | ···················· | ··A················· |
| F | 1469 hCoV-19/Bangladesh/BCSIR-NILMRC-382/2020\|EPI_ISL_5 | ···················· | ···················· | ···················· | ···················· | ···················· | ···················· |
| F | 1470 hCoV-19/Bangladesh/BCSIR-NILMRC-371/2020\|EPI_ISL_5 | ···················· | ···················· | ···················· | ···················· | ···················· | ···················· |
| F | 1471 hCoV-19/Bangladesh/BCSIR-NILMRC-372/2020\|EPI_ISL_5 | ···················· | ···················· | ···················· | ···················· | ···················· | ···················· |
| F | 1472 hCoV-19/Bangladesh/BCSIR-NILMRC-374/2020\|EPI_ISL_5 | ···················· | ···················· | ···················· | ···················· | ···················· | ···················· |
| F | 1473 hCoV-19/Bangladesh/BCSIR-NILMRC-383/2020\|EPI_ISL_5 | ···················· | ···················· | ···················· | ···················· | ···················· | ···················· |
| F | 1474 hCoV-19/Bangladesh/BCSIR-NILMRC-384/2020\|EPI_ISL_5 | ···················· | ···················· | ···················· | ···················· | ···················· | ···················· |
| F | 1475 hCoV-19/Bangladesh/BCSIR-DU-16/2020\|EPI_ISL_514253 | ···················· | ·A·················· | ···················· | ···················· | ···················· | ···················· |
| F | 1476 hCoV-19/Peru/LAM-INS-043/2020\|EPI_ISL_514264\|2020- | ···················· | ···················· | ···················· | ···················· | ···················· | ···················· |
| F | 1477 hCoV-19/Peru/LAM-INS-044/2020\|EPI_ISL_514265\|2020- | ···················· | ···················· | ···················· | ···················· | ···················· | ···················· |
| F | 1478 hCoV-19/Israel/CVL-n25334/2020\|EPI_ISL_514266\|2020 | ···················· | ···················· | ···················· | ···················· | ···················· | ···················· |
| F | 1479 hCoV-19/Israel/CVL-n25794/2020\|EPI_ISL_514268\|2020 | ···················· | ·A·················· | ···················· | ···················· | ···················· | ···················· |
| F | 1480 hCoV-19/Israel/CVL-n25863/2020\|EPI_ISL_514269\|2020 | ···················· | ···················· | ···················· | ···················· | ···················· | ···················· |
| F | 1481 hCoV-19/Israel/CVL-n25867/2020\|EPI_ISL_514270\|2020 | ···················· | ···················· | ··········T········· | ···················· | ···················· | ···················· |
| F | 1483 hCoV-19/Israel/CVL-n25872/2020\|EPI_ISL_514272\|2020 | ···················· | ·A·················· | ···················· | ···················· | ···················· | ···················· |
| F | 1484 hCoV-19/Israel/CVL-n25886/2020\|EPI_ISL_514274\|2020 | ···················· | ·A·················· | ···················· | ···················· | ···················· | ···················· |
| F | 1485 hCoV-19/Israel/CVL-n25904/2020\|EPI_ISL_514275\|2020 | ···················· | ···················· | ···················· | ···················· | ···················· | ···················· |
| F | 1487 hCoV-19/Israel/CVL-n25924/2020\|EPI_ISL_514279\|2020 | ···················· | ···················· | ···················· | ···················· | ···················· | ···················· |
| F | 1488 hCoV-19/Israel/CVL-n25929/2020\|EPI_ISL_514280\|2020 | ···················· | ·A·················· | ···················· | ···················· | ···················· | ···················· |
| F | 1489 hCoV-19/Israel/CVL-n25947/2020\|EPI_ISL_514281\|2020 | ···················· | ·A·················· | ···················· | ···················· | ···················· | ···················· |
| F | 1490 hCoV-19/Israel/CVL-n26099/2020\|EPI_ISL_514283\|2020 | ···················· | ···················· | ···················· | ···················· | ···················· | ···················· |
| F | 1491 hCoV-19/Israel/CVL-n26111/2020\|EPI_ISL_514284\|2020 | ···················· | ···················· | ···················· | ···················· | ···················· | ···················· |
| F | 1492 hCoV-19/Israel/CVL-n26113/2020\|EPI_ISL_514286\|2020 | ···················· | ···················· | ···················· | ···················· | ···················· | ···················· |
| F | 1493 hCoV-19/Israel/CVL-n26122/2020\|EPI_ISL_514287\|2020 | ···················· | ···················· | ···················· | ···················· | ···················· | ···················· |
| F | 1494 hCoV-19/Israel/CVL-n26131/2020\|EPI_ISL_514288\|2020 | ···················· | ···················· | ··········T········· | ···················· | ···················· | ···················· |
| F | 1495 hCoV-19/Israel/CVL-n26134/2020\|EPI_ISL_514289\|2020 | ···················· | ···················· | ···················· | ···················· | ···················· | ···················· |
| F | 1496 hCoV-19/Brazil/SP-627/2020\|EPI_ISL_515541\|2020-03- | ···················· | ···················· | ···················· | ···················· | ···················· | ···················· |
| F | 1497 hCoV-19/Brazil/SP-136/2020\|EPI_ISL_515542\|2020-04- | ···················· | ···················· | ···················· | ···················· | ···················· | ···················· |
| F | 1498 hCoV-19/Brazil/SP-137/2020\|EPI_ISL_515543\|2020-04- | ···················· | ···················· | ···················· | ···················· | ···················· | ···················· |
| F | 1499 hCoV-19/Brazil/SP-629/2020\|EPI_ISL_515544\|2020-03- | ···················· | ···················· | ···················· | ···················· | ···················· | ···················· |
| F | 1500 hCoV-19/Brazil/SP-533/2020\|EPI_ISL_515545\|2020-03- | ···················· | ···················· | ···················· | ···················· | ···················· | ···················· |
| F | 1501 hCoV-19/Brazil/SP-632/2020\|EPI_ISL_515546\|2020-03- | ···················· | ···················· | ···················· | ···················· | ···················· | ···················· |
| F | 1502 hCoV-19/Brazil/SP-633/2020\|EPI_ISL_515547\|2020-03- | ···················· | ···················· | ···················· | ···················· | ···················· | ···················· |
| F | 1503 hCoV-19/Brazil/SP-634/2020\|EPI_ISL_515548\|2020-03- | ···················· | ···················· | ···················· | ···················· | ···················· | ···················· |
| F | 1504 hCoV-19/Brazil/SP-636/2020\|EPI_ISL_515550\|2020-03- | ···················· | ···················· | ···················· | ···················· | ···················· | ···················· |
| F | 1505 hCoV-19/Brazil/SP-544/2020\|EPI_ISL_515551\|2020-03- | ···················· | ···················· | ···················· | ···················· | ···················· | ···················· |
| F | 1506 hCoV-19/Brazil/SP-611/2020\|EPI_ISL_515552\|2020-04- | ···················· | ···················· | ···················· | ···················· | ···················· | ···················· |
| F | 1507 hCoV-19/Brazil/SP-612/2020\|EPI_ISL_515553\|2020-04- | ···················· | ···················· | ···················· | ···················· | ···················· | ···················· |
| F | 1508 hCoV-19/Brazil/SP-639/2020\|EPI_ISL_515554\|2020-03- | ···················· | ···················· | ···················· | ···················· | ···················· | ···················· |
| F | 1509 hCoV-19/Brazil/SP-154/2020\|EPI_ISL_515555\|2020-03- | ···················· | ···················· | ···················· | ···················· | ···················· | ···················· |
| F | 1510 hCoV-19/Brazil/SP-615/2020\|EPI_ISL_515557\|2020-04- | ···················· | ···················· | ···················· | ···················· | ···················· | ···················· |
| F | 1511 hCoV-19/Brazil/SP-155/2020\|EPI_ISL_515559\|2020-04- | ···················· | ···················· | ···················· | ···················· | ···················· | ···················· |
| F | 1512 hCoV-19/Brazil/SP-156/2020\|EPI_ISL_515560\|2020-04- | ···················· | ···················· | ···················· | ···················· | ···················· | ···················· |
| F | 1513 hCoV-19/Brazil/SP-617/2020\|EPI_ISL_515561\|2020-04- | ···················· | ···················· | ···················· | ···················· | ···················· | ···················· |
| F | 1514 hCoV-19/Brazil/SP-619/2020\|EPI_ISL_515562\|2020-04- | ···················· | ···················· | ···················· | ···················· | ···················· | ···················· |
| F | 1515 hCoV-19/Brazil/SP-159/2020\|EPI_ISL_515563\|2020-04- | ···················· | ···················· | ···················· | ···················· | ···················· | ···················· |
| F | 1516 hCoV-19/Brazil/SP-187/2020\|EPI_ISL_515564\|2020-03- | ···················· | ···················· | ···················· | ···················· | ···················· | ···················· |

| | | | | | | | | |
|---|---|---|---|---|---|---|---|---|
| F | 1517 | hCoV-19/Brazil/SP-188/2020\|EPI_ISL_515565\|2020-03- | .................... | .................... | .................... | .................... | .................... | .................... |
| F | 1518 | hCoV-19/Brazil/SP-623/2020\|EPI_ISL_515566\|2020-04- | .................... | .................... | .................... | .................... | .................... | .................... |
| F | 1519 | hCoV-19/South Africa/KRISP-K002162/2020\|EPI_ISL_51 | .................... | .................... | .................... | .................... | .................... | .................... |
| F | 1520 | hCoV-19/South Africa/KRISP-K002163/2020\|EPI_ISL_51 | .................... | .................... | .................... | .................... | .................... | .................... |
| F | 1521 | hCoV-19/South Africa/KRISP-K002164/2020\|EPI_ISL_51 | .................... | .................... | .................... | .................... | .................... | .................... |
| F | 1522 | hCoV-19/South Africa/KRISP-K002171/2020\|EPI_ISL_51 | .................... | .................... | .................... | .................... | .................... | .................... |
| F | 1523 | hCoV-19/South Africa/KRISP-K002172/2020\|EPI_ISL_51 | .................... | .................... | .................... | .................... | .................... | .................... |
| F | 1524 | hCoV-19/South Africa/KRISP-K002173/2020\|EPI_ISL_51 | .................... | .................... | .................... | .................... | .................... | .................... |
| F | 1525 | hCoV-19/South Africa/KRISP-K002177/2020\|EPI_ISL_51 | .................... | .................... | .................... | .................... | .................... | .................... |
| F | 1526 | hCoV-19/South Africa/KRISP-K002179/2020\|EPI_ISL_51 | .................... | .................... | .................... | .................... | .................... | .................... |
| F | 1527 | hCoV-19/South Africa/KRISP-K002183/2020\|EPI_ISL_51 | .................... | .................... | .................... | .................... | .................... | .................... |
| F | 1528 | hCoV-19/South Africa/KRISP-K002185/2020\|EPI_ISL_51 | .................... | .................... | .................... | .................... | .................... | .................... |
| F | 1529 | hCoV-19/South Africa/KRISP-K002186/2020\|EPI_ISL_51 | .................... | .................... | .................... | .................... | .................... | .................... |
| F | 1530 | hCoV-19/South Africa/KRISP-K002187/2020\|EPI_ISL_51 | .................... | .................... | .................... | .................... | .................... | .................... |
| F | 1531 | hCoV-19/South Africa/KRISP-K002190/2020\|EPI_ISL_51 | .................... | .................... | .................... | .................... | .................... | .................... |
| F | 1532 | hCoV-19/South Africa/KRISP-K002194/2020\|EPI_ISL_51 | .................... | .................... | .................... | .................... | .................... | .................... |
| F | 1533 | hCoV-19/South Africa/KRISP-K002195/2020\|EPI_ISL_51 | .................... | .................... | .................... | .................... | .................... | .................... |
| F | 1534 | hCoV-19/South Africa/KRISP-K002196/2020\|EPI_ISL_51 | .................... | .................... | .................... | .................... | .................... | .................... |
| F | 1535 | hCoV-19/South Africa/KRISP-K002198/2020\|EPI_ISL_51 | .................... | .................... | .................... | .................... | .................... | .................... |
| F | 1536 | hCoV-19/South Africa/KRISP-K002199/2020\|EPI_ISL_51 | .................... | .................... | .................... | .................... | .................... | .................... |
| F | 1537 | hCoV-19/USA/WA-S2416/2020\|EPI_ISL_497048\|2020-06-1 | .................... | .................... | .................... | .................... | .................... | .................... |
| F | 1538 | hCoV-19/USA/WA-S2417/2020\|EPI_ISL_497049\|2020-06-1 | .................... | .................... | .................... | .................... | .................... | .................... |
| F | 1539 | hCoV-19/USA/WA-S2418/2020\|EPI_ISL_497050\|2020-06-1 | .................... | .................... | .................... | .................... | .................... | .................... |
| F | 1540 | hCoV-19/USA/WA-S2419/2020\|EPI_ISL_497051\|2020-06-1 | .................... | ....A............... | .................... | .................... | .................... | .................... |
| F | 1541 | hCoV-19/USA/WA-S2420/2020\|EPI_ISL_497052\|2020-06-1 | .................... | .................... | .................... | .................... | .................... | .................... |
| F | 1542 | hCoV-19/USA/WA-S2422/2020\|EPI_ISL_497054\|2020-06-1 | .................... | .................... | .................... | .................... | .................... | .................... |
| F | 1543 | hCoV-19/USA/WA-S2423/2020\|EPI_ISL_497055\|2020-06-1 | .................... | .................... | .................... | .................... | .................... | .................... |
| F | 1544 | hCoV-19/USA/WA-S2424/2020\|EPI_ISL_497056\|2020-06-1 | .................... | .................... | .................... | .................... | .................... | .................... |
| F | 1545 | hCoV-19/USA/WA-S2425/2020\|EPI_ISL_497057\|2020-06-1 | .................... | .................... | .................... | .................... | .................... | .................... |
| F | 1546 | hCoV-19/USA/WA-S2426/2020\|EPI_ISL_497058\|2020-06-1 | .................... | .................... | .................... | .................... | .................... | .................... |
| F | 1547 | hCoV-19/USA/WA-S2427/2020\|EPI_ISL_497059\|2020-06-1 | .................... | .................... | .................... | .................... | .................... | .................... |
| F | 1548 | hCoV-19/USA/WA-S2428/2020\|EPI_ISL_497060\|2020-06-1 | .................... | .................... | .................... | .................... | .................... | .................... |
| F | 1549 | hCoV-19/USA/WA-S2429/2020\|EPI_ISL_497061\|2020-06-1 | .................... | .................... | .................... | .................... | .................... | .................... |
| F | 1550 | hCoV-19/USA/WA-S2430/2020\|EPI_ISL_497062\|2020-06-1 | .................... | .................... | .................... | .................... | .................... | .................... |
| F | 1551 | hCoV-19/USA/WA-S2431/2020\|EPI_ISL_497063\|2020-06-1 | .................... | .................... | .................... | .................... | .................... | .................... |
| F | 1552 | hCoV-19/USA/WA-S2432/2020\|EPI_ISL_497064\|2020-06-1 | .................... | .................... | .................... | .................... | .................... | .................... |
| F | 1553 | hCoV-19/USA/WA-S2433/2020\|EPI_ISL_497065\|2020-06-1 | .................... | .................... | .................... | .................... | .................... | .................... |
| F | 1554 | hCoV-19/USA/WA-S2434/2020\|EPI_ISL_497066\|2020-06-1 | .................... | .................... | .................... | .................... | .................... | .................... |
| F | 1555 | hCoV-19/USA/WA-S2435/2020\|EPI_ISL_497067\|2020-06-1 | .................... | .................... | .................... | .................... | .................... | .................... |
| F | 1556 | hCoV-19/USA/WA-S2436/2020\|EPI_ISL_497068\|2020-06-1 | .................... | .................... | .................... | .................... | .................... | .................... |
| F | 1557 | hCoV-19/USA/WA-S2437/2020\|EPI_ISL_497069\|2020-06-1 | .................... | .................... | .................... | .................... | .................... | .................... |
| F | 1558 | hCoV-19/USA/WA-S2438/2020\|EPI_ISL_497070\|2020-06-1 | .................... | .................... | .................... | .................... | .................... | .................... |
| F | 1559 | hCoV-19/USA/WA-S2439/2020\|EPI_ISL_497071\|2020-06-1 | .................... | .................... | .................... | .................... | .................... | .................... |
| F | 1560 | hCoV-19/USA/WA-S2440/2020\|EPI_ISL_497072\|2020-06-1 | .................... | .................... | .................... | .................... | .................... | .................... |
| F | 1561 | hCoV-19/USA/WA-S2442/2020\|EPI_ISL_497074\|2020-06-1 | .................... | .................... | .................... | .................... | .................... | .................... |
| F | 1562 | hCoV-19/USA/WA-S2443/2020\|EPI_ISL_497075\|2020-06-1 | .................... | .................... | .................... | .................... | .................... | .................... |
| F | 1563 | hCoV-19/USA/WA-S2444/2020\|EPI_ISL_497076\|2020-06-1 | .................... | .................... | .................... | .................... | .................... | .................... |
| F | 1564 | hCoV-19/USA/WA-S2448/2020\|EPI_ISL_497080\|2020-06-1 | .................... | .................... | .................... | .................... | .................... | .................... |
| F | 1565 | hCoV-19/USA/WA-S2449/2020\|EPI_ISL_497081\|2020-06-1 | .................... | .................... | .................... | .................... | .................... | .................... |
| F | 1566 | hCoV-19/USA/WA-S2450/2020\|EPI_ISL_497082\|2020-06-1 | .................... | .................... | .................... | .................... | .................... | .................... |
| F | 1567 | hCoV-19/USA/WA-S2451/2020\|EPI_ISL_497083\|2020-06-1 | .................... | .................... | .................... | .................... | .................... | .................... |
| F | 1568 | hCoV-19/USA/WA-S2452/2020\|EPI_ISL_497084\|2020-06-1 | .................... | .................... | .................... | .................... | .................... | .................... |
| F | 1569 | hCoV-19/USA/WA-S2453/2020\|EPI_ISL_497085\|2020-06-1 | .................... | .................... | .................... | .................... | .................... | .................... |
| F | 1570 | hCoV-19/USA/WA-S2454/2020\|EPI_ISL_497086\|2020-06-1 | .................... | .................... | .................... | .................... | .................... | .................... |
| F | 1571 | hCoV-19/USA/WA-S2455/2020\|EPI_ISL_497087\|2020-06-1 | .................... | .................... | .................... | .................... | .................... | .................... |
| F | 1572 | hCoV-19/USA/WA-S2456/2020\|EPI_ISL_497088\|2020-06-1 | .................... | .................... | .................... | .................... | .................... | .................... |
| F | 1573 | hCoV-19/USA/WA-S2457/2020\|EPI_ISL_497089\|2020-06-1 | .................... | .................... | .................... | .................... | .................... | .................... |
| F | 1574 | hCoV-19/England/NORT-29A5EC/2020\|EPI_ISL_507114\|20 | .................... | .................... | .................... | .................... | .................... | .................... |
| F | 1575 | hCoV-19/India/MH-GA55/2020\|EPI_ISL_508250\|2020-06- | .................... | .................... | .................... | .................... | .................... | .................... |
| F | 1576 | hCoV-19/India/MH-GA56/2020\|EPI_ISL_508251\|2020-06- | .................... | .................... | .................... | .................... | .................... | .................... |
| F | 1577 | hCoV-19/India/MH-GA57/2020\|EPI_ISL_508252\|2020-06- | .................... | .................... | .................... | .................... | .................... | .................... |
| F | 1578 | hCoV-19/India/MH-GA59/2020\|EPI_ISL_508253\|2020-06- | .................... | .................... | .................... | .................... | .................... | .................... |
| F | 1579 | hCoV-19/India/MH-GA6/2020\|EPI_ISL_508254\|2020-06-1 | .................... | .................... | .................... | .................... | .................... | .................... |
| F | 1580 | hCoV-19/India/MH-GA60/2020\|EPI_ISL_508255\|2020-06- | .................... | .................... | .................... | .................... | .................... | .................... |
| F | 1581 | hCoV-19/India/MH-GA61/2020\|EPI_ISL_508256\|2020-06- | .................... | .................... | .................... | .................... | .................... | .................... |
| F | 1582 | hCoV-19/India/MH-GA62/2020\|EPI_ISL_508257\|2020-06- | .................... | .................... | .................... | .................... | .................... | .................... |
| F | 1583 | hCoV-19/India/MH-GA63/2020\|EPI_ISL_508258\|2020-06- | .................... | .................... | .................... | .................... | .................... | .................... |
| F | 1584 | hCoV-19/India/MH-GA65/2020\|EPI_ISL_508259\|2020-06- | .................... | .................... | .................... | .................... | .................... | .................... |
| F | 1585 | hCoV-19/India/MH-GA66/2020\|EPI_ISL_508260\|2020-06- | .................... | .................... | .................... | .................... | .................... | .................... |
| F | 1586 | hCoV-19/India/MH-GA67/2020\|EPI_ISL_508261\|2020-06- | .................... | .................... | .................... | .................... | .................... | .................... |
| F | 1587 | hCoV-19/India/MH-GA68/2020\|EPI_ISL_508262\|2020-06- | .................... | .................... | .................... | .................... | .................... | .................... |
| F | 1588 | hCoV-19/India/MH-GA69/2020\|EPI_ISL_508263\|2020-06- | .................... | .................... | .................... | .................... | .................... | .................... |
| F | 1589 | hCoV-19/India/MH-GA7/2020\|EPI_ISL_508264\|2020-06-1 | .................... | .................... | .................... | .................... | .................... | .................... |
| F | 1590 | hCoV-19/India/MH-GA70/2020\|EPI_ISL_508265\|2020-06- | .................... | .................... | .................... | .................... | .................... | .................... |
| F | 1591 | hCoV-19/India/MH-GA71/2020\|EPI_ISL_508266\|2020-06- | .................... | .................... | .................... | .................... | .................... | .................... |
| F | 1592 | hCoV-19/India/MH-GA73/2020\|EPI_ISL_508267\|2020-06- | .................... | .................... | .................... | .................... | .................... | .................... |
| F | 1593 | hCoV-19/India/MH-GA74/2020\|EPI_ISL_508268\|2020-06- | .................... | .................... | .................... | .................... | .................... | .................... |
| F | 1594 | hCoV-19/India/MH-GA75/2020\|EPI_ISL_508269\|2020-06- | .................... | .................... | .................... | .................... | .................... | .................... |
| F | 1595 | hCoV-19/India/MH-GA76/2020\|EPI_ISL_508270\|2020-06- | .................... | .................... | .................... | .................... | .................... | .................... |
| F | 1596 | hCoV-19/India/MH-GA77/2020\|EPI_ISL_508271\|2020-06- | .................... | .................... | .................... | .................... | .................... | .................... |

| | | | | | | | | |
|---|---|---|---|---|---|---|---|---|
| F | 1597 hCoV-19/India/MH-GA78/2020\|EPI_ISL_508272\|2020-06- | ···················· | ···················· | ···················· | ···················· | ···················· | ···················· | |
| F | 1598 hCoV-19/India/MH-GA79/2020\|EPI_ISL_508273\|2020-06- | ···················· | ···················· | ···················· | ···················· | ···················· | ···················· | |
| F | 1599 hCoV-19/India/MH-GA8/2020\|EPI_ISL_508274\|2020-06-1 | ···················· | ···················· | ···················· | ···················· | ···················· | ···················· | |
| F | 1600 hCoV-19/India/MH-GA80/2020\|EPI_ISL_508275\|2020-06- | ···················· | ···················· | ···················· | ···················· | ···················· | ···················· | |
| F | 1601 hCoV-19/India/MH-GA81/2020\|EPI_ISL_508276\|2020-06- | ···················· | ···················· | ···················· | ···················· | ···················· | ···················· | |
| F | 1602 hCoV-19/India/MH-GA82/2020\|EPI_ISL_508277\|2020-06- | ···················· | ···················· | ···················· | ···················· | ···················· | ···················· | |
| F | 1603 hCoV-19/India/MH-GA83/2020\|EPI_ISL_508278\|2020-06- | ···················· | ···················· | ···················· | ···················· | ···················· | ···················· | |
| F | 1604 hCoV-19/India/MH-GA84/2020\|EPI_ISL_508279\|2020-06- | ···················· | ···················· | ···················· | ···················· | ···················· | ···················· | |
| F | 1605 hCoV-19/India/MH-GA85/2020\|EPI_ISL_508280\|2020-06- | ···················· | ···················· | ···················· | ···················· | ···················· | ···················· | |
| F | 1606 hCoV-19/India/MH-GA86/2020\|EPI_ISL_508281\|2020-06- | ···················· | ···················· | ···················· | ···················· | ···················· | ···················· | |
| F | 1607 hCoV-19/India/MH-GA87/2020\|EPI_ISL_508282\|2020-06- | ···················· | ···················· | ···················· | ···················· | ···················· | ···················· | |
| F | 1608 hCoV-19/India/MH-GA88/2020\|EPI_ISL_508283\|2020-06- | ···················· | ···················· | ···················· | ···················· | ···················· | ···················· | |
| F | 1609 hCoV-19/India/MH-GA9/2020\|EPI_ISL_508284\|2020-06-1 | ···················· | ···················· | ···················· | ···················· | ···················· | ···················· | |
| F | 1610 hCoV-19/India/MH-GA93/2020\|EPI_ISL_508285\|2020-06- | ···················· | ···················· | ···················· | ···················· | ···················· | ···················· | |
| F | 1611 hCoV-19/India/MH-GA98/2020\|EPI_ISL_508286\|2020-06- | ···················· | ···················· | ···················· | ···················· | ···················· | ···················· | |
| F | 1612 hCoV-19/India/KA-IB1/2020\|EPI_ISL_508287\|2020-06-1 | ···················· | ···················· | ···················· | ···················· | ···················· | ···················· | |
| F | 1613 hCoV-19/India/KA-IB10/2020\|EPI_ISL_508288\|2020-06- | ···················· | ···················· | ···················· | ···················· | ···················· | ···················· | |
| F | 1614 hCoV-19/India/KA-IB11/2020\|EPI_ISL_508289\|2020-06- | ···················· | ···················· | ···················· | ···················· | ···················· | ···················· | |
| F | 1615 hCoV-19/India/WB-IK79/2020\|EPI_ISL_508400\|2020-05- | ···················· | ···················· | ···················· | ···················· | ···················· | ···················· | |
| F | 1616 hCoV-19/India/WB-IK80/2020\|EPI_ISL_508401\|2020-05- | ···················· | ···················· | ···················· | ···················· | ···················· | ···················· | |
| F | 1617 hCoV-19/India/WB-IK82/2020\|EPI_ISL_508402\|2020-05- | ···················· | ···················· | ···················· | ···················· | ···················· | ···················· | |
| F | 1618 hCoV-19/India/WB-IK83/2020\|EPI_ISL_508403\|2020-05- | ···················· | ···················· | ···················· | ···················· | ···················· | ···················· | |
| F | 1619 hCoV-19/India/WB-IK85/2020\|EPI_ISL_508404\|2020-05- | ···················· | ···················· | ···················· | ···················· | ···················· | ···················· | |
| F | 1620 hCoV-19/India/WB-IK86/2020\|EPI_ISL_508405\|2020-05- | ···················· | ···················· | ···················· | ···················· | ···················· | ···················· | |
| F | 1621 hCoV-19/India/WB-IK88/2020\|EPI_ISL_508406\|2020-05- | ···················· | ···················· | ···················· | ···················· | ···················· | ···················· | |
| F | 1622 hCoV-19/India/WB-IK9/2020\|EPI_ISL_508407\|2020-04-1 | ···················· | ···················· | ···················· | ···················· | ···················· | ···················· | |
| F | 1623 hCoV-19/India/WB-IK90/2020\|EPI_ISL_508408\|2020-05- | ···················· | ···················· | ···················· | ···················· | ···················· | ···················· | |
| F | 1624 hCoV-19/India/WB-IK91/2020\|EPI_ISL_508409\|2020-05- | ···················· | ···················· | ···················· | ···················· | ···················· | ···················· | |
| F | 1625 hCoV-19/India/WB-IK92/2020\|EPI_ISL_508410\|2020-05- | ···················· | ···················· | ···················· | ···················· | ·················C·· | ···················· | 1 |
| F | 1626 hCoV-19/India/WB-IK93/2020\|EPI_ISL_508411\|2020-05- | ···················· | ···················· | ···················· | ···················· | ··A················· | ···················· | |
| F | 1627 hCoV-19/India/WB-IK96/2020\|EPI_ISL_508412\|2020-05- | ···················· | ···················· | ···················· | ···················· | ···················· | ···················· | |
| F | 1628 hCoV-19/India/WB-IK97/2020\|EPI_ISL_508413\|2020-05- | ···················· | ···················· | ···················· | ···················· | ···················· | ···················· | |
| F | 1629 hCoV-19/India/WB-IK99/2020\|EPI_ISL_508414\|2020-05- | ···················· | ···················· | ···················· | ···················· | ···················· | ···················· | |
| F | 1630 hCoV-19/India/DL-MN1/2020\|EPI_ISL_508415\|2020-06-1 | ···················· | ·A·················· | ···················· | ···················· | ···················· | ··A················· | |
| F | 1631 hCoV-19/India/DL-MN10/2020\|EPI_ISL_508416\|2020-06- | ···················· | ···················· | ···················· | ···················· | ···················· | ···················· | |
| F | 1632 hCoV-19/India/DL-MN11/2020\|EPI_ISL_508417\|2020-06- | ···················· | ···················· | ···················· | ···················· | ···················· | ···················· | |
| F | 1633 hCoV-19/India/DL-MN26/2020\|EPI_ISL_508418\|2020-06- | ···················· | ·A·················· | ···················· | ···················· | ···················· | ··A················· | |
| F | 1634 hCoV-19/India/UP-MN32/2020\|EPI_ISL_508419\|2020-06- | ···················· | ·A·················· | ···················· | ···················· | ···················· | ··A················· | |
| F | 1635 hCoV-19/India/DL-MN4/2020\|EPI_ISL_508420\|2020-06-1 | ···················· | ·A·················· | ···················· | ···················· | ···················· | ··A················· | |
| F | 1636 hCoV-19/India/DL-MN5/2020\|EPI_ISL_508421\|2020-06-1 | ···················· | ···················· | ···················· | ···················· | ···················· | ···················· | |
| F | 1637 hCoV-19/India/DL-MN8/2020\|EPI_ISL_508422\|2020-06-1 | ···················· | ·A·················· | ···················· | ···················· | ···················· | ··A················· | |
| F | 1638 hCoV-19/India/MH-MW11/2020\|EPI_ISL_508423\|2020-04- | ···················· | ···················· | ···················· | ···················· | ···················· | ···················· | |
| F | 1639 hCoV-19/India/MH-MW12/2020\|EPI_ISL_508424\|2020-05- | ···················· | ···················· | ···················· | ···················· | ···················· | ···················· | |
| F | 1640 hCoV-19/India/MH-MW17/2020\|EPI_ISL_508425\|2020-05- | ···················· | ···················· | ···················· | ···················· | ···················· | ···················· | |
| F | 1641 hCoV-19/India/MH-MW18/2020\|EPI_ISL_508426\|2020-05- | ···················· | ···················· | ···················· | ···················· | ···················· | ···················· | |
| F | 1642 hCoV-19/India/MH-MW20/2020\|EPI_ISL_508427\|2020-05- | ···················· | ···················· | ···················· | ···················· | ···················· | ···················· | |
| F | 1643 hCoV-19/India/UP-MW24/2020\|EPI_ISL_508428\|2020-05- | ···················· | ···················· | ···················· | ···················· | ···················· | ···················· | |
| F | 1644 hCoV-19/India/MH-MW25/2020\|EPI_ISL_508429\|2020-05- | ···················· | ···················· | ···················· | ···················· | ···················· | ···················· | |
| F | 1645 hCoV-19/India/WB-S36/2020\|EPI_ISL_508445\|2020-05-0 | ···················· | ···················· | ···················· | ···················· | ···················· | ···················· | |
| F | 1646 hCoV-19/USA/UT-03739/2020\|EPI_ISL_509580\|2020-04-0 | ···················· | ···················· | ···················· | ···················· | ···················· | ···················· | |
| F | 1647 hCoV-19/USA/UT-03769/2020\|EPI_ISL_509585\|2020-04-0 | ···················· | ···················· | ···················· | ···················· | ···················· | ···················· | |
| F | 1648 hCoV-19/USA/UT-03777/2020\|EPI_ISL_509588\|2020-04-0 | ···················· | ···················· | ···················· | ···················· | ···················· | ···················· | |
| F | 1649 hCoV-19/USA/UT-03778/2020\|EPI_ISL_509589\|2020-04-0 | ···················· | ···················· | ···················· | ···················· | ···················· | ···················· | |
| F | 1650 hCoV-19/USA/UT-03780/2020\|EPI_ISL_509590\|2020-04-0 | ···················· | ···················· | ···················· | ···················· | ···················· | ···················· | |
| F | 1651 hCoV-19/USA/UT-03784/2020\|EPI_ISL_509592\|2020-04-0 | ···················· | ···················· | ···················· | ···················· | ···················· | ···················· | |
| F | 1652 hCoV-19/Guatemala/CDC-1233/2020\|EPI_ISL_509700\|202 | ···················· | ···················· | ···················· | ···················· | ···················· | ···················· | |
| F | 1653 hCoV-19/Guatemala/CDC-1234/2020\|EPI_ISL_509701\|202 | ···················· | ···················· | ···················· | ···················· | ···················· | ···················· | |
| F | 1654 hCoV-19/Guatemala/CDC-1235/2020\|EPI_ISL_509702\|202 | ···················· | ···················· | ···················· | ···················· | ···················· | ···················· | |
| F | 1655 hCoV-19/Guatemala/CDC-1236/2020\|EPI_ISL_509703\|202 | ···················· | ···················· | ···················· | ···················· | ···················· | ···················· | |
| F | 1656 hCoV-19/USA/WI-CDC-0944/2020\|EPI_ISL_509704\|2020-0 | ···················· | ···················· | ···················· | ···················· | ···················· | ···················· | |
| F | 1657 hCoV-19/USA/WI-CDC-0946/2020\|EPI_ISL_509705\|2020-0 | ···················· | ···················· | ···················· | ···················· | ···················· | ···················· | |
| F | 1658 hCoV-19/USA/WI-CDC-0953/2020\|EPI_ISL_509706\|2020-0 | ···················· | ···················· | ···················· | ···················· | ···················· | ···················· | |
| F | 1659 hCoV-19/USA/UT-CDC-9770/2020\|EPI_ISL_509707\|2020-0 | ···················· | ···················· | ···················· | ···················· | ···················· | ···················· | |
| F | 1660 hCoV-19/USA/UT-CDC-9774/2020\|EPI_ISL_509708\|2020-0 | ···················· | ···················· | ···················· | ···················· | ···················· | ···················· | |
| F | 1661 hCoV-19/USA/UT-CDC-9782/2020\|EPI_ISL_509709\|2020-0 | ···················· | ···················· | ···················· | ···················· | ···················· | ···················· | |
| F | 1662 hCoV-19/Guatemala/CDC-1229/2020\|EPI_ISL_509710\|202 | ···················· | ···················· | ···················· | ···················· | ···················· | ···················· | |
| F | 1663 hCoV-19/Belize/CDC-6840/2020\|EPI_ISL_509711\|2020-0 | ···················· | ···················· | ···················· | ···················· | ···················· | ···················· | |
| F | 1664 hCoV-19/Belize/CDC-6846/2020\|EPI_ISL_509712\|2020-0 | ···················· | ···················· | ···················· | ···················· | ···················· | ···················· | |
| F | 1665 hCoV-19/Belize/CDC-6851/2020\|EPI_ISL_509713\|2020-0 | ···················· | ···················· | ···················· | ···················· | ···················· | ···················· | |
| F | 1666 hCoV-19/Belize/CDC-6852/2020\|EPI_ISL_509714\|2020-0 | ···················· | ···················· | ···················· | ···················· | ···················· | ···················· | |
| F | 1667 hCoV-19/USA/WI-CDC-0952/2020\|EPI_ISL_509715\|2020-0 | ···················· | ···················· | ···················· | ···················· | ···················· | ···················· | |
| F | 1668 hCoV-19/USA/FL-BPHL-0658/2020\|EPI_ISL_509716\|2020- | ···················· | ···················· | ···················· | ···················· | ···················· | ···················· | |
| F | 1669 hCoV-19/USA/FL-BPHL-0659/2020\|EPI_ISL_509717\|2020- | ···················· | ···················· | ···················· | ···················· | ···················· | ···················· | |
| F | 1670 hCoV-19/USA/FL-BPHL-0660/2020\|EPI_ISL_509718\|2020- | ···················· | ···················· | ···················· | ···················· | ···················· | ···················· | |
| F | 1671 hCoV-19/USA/FL-BPHL-0661/2020\|EPI_ISL_509719\|2020- | ···················· | ···················· | ···················· | ···················· | ···················· | ···················· | |
| F | 1672 hCoV-19/USA/FL-BPHL-0663/2020\|EPI_ISL_509721\|2020- | ···················· | ···················· | ···················· | ···················· | ···················· | ···················· | |
| F | 1673 hCoV-19/USA/FL-BPHL-0664/2020\|EPI_ISL_509722\|2020- | ···················· | ···················· | ···················· | ···················· | ···················· | ···················· | |
| F | 1675 hCoV-19/USA/FL-BPHL-0666/2020\|EPI_ISL_509724\|2020- | ···················· | ···················· | ···················· | ···················· | ···················· | ···················· | |
| F | 1676 hCoV-19/USA/FL-BPHL-0667/2020\|EPI_ISL_509725\|2020- | ···················· | ···················· | ···················· | ···················· | ···················· | ···················· | |
| F | 1677 hCoV-19/USA/FL-BPHL-0668/2020\|EPI_ISL_509726\|2020- | ···················· | ···················· | ···················· | ···················· | ···················· | ···················· | |

| | | | | | | | | |
|---|---|---|---|---|---|---|---|---|
| F | 1678 | hCoV-19/USA/FL-BPHL-0669/2020\|EPI_ISL_509727\|2020- | ···················· | ···················· | ···················· | ···················· | ···················· | ···················· |
| F | 1679 | hCoV-19/USA/FL-BPHL-0670/2020\|EPI_ISL_509728\|2020- | ···················· | ···················· | ···················· | ···················· | ···················· | ···················· |
| F | 1681 | hCoV-19/USA/FL-BPHL-0672/2020\|EPI_ISL_509730\|2020- | ···················· | ···················· | ···················· | ···················· | ···················· | ···················· |
| F | 1682 | hCoV-19/USA/FL-BPHL-0673/2020\|EPI_ISL_509731\|2020- | ···················· | ···················· | ···················· | ···················· | ···················· | ···················· |
| F | 1683 | hCoV-19/USA/FL-BPHL-0674/2020\|EPI_ISL_509732\|2020- | ···················· | ···················· | ···················· | ···················· | ···················· | ···················· |
| F | 1685 | hCoV-19/USA/FL-BPHL-0676/2020\|EPI_ISL_509734\|2020- | ···················· | ···················· | ···················· | ···················· | ···················· | ···················· |
| F | 1686 | hCoV-19/USA/FL-BPHL-0677/2020\|EPI_ISL_509735\|2020- | ···················· | ···················· | ···················· | ···················· | ···················· | ···················· |
| F | 1687 | hCoV-19/USA/FL-BPHL-0678/2020\|EPI_ISL_509736\|2020- | ···················· | ···················· | ···················· | ···················· | ···················· | ···················· |
| F | 1689 | hCoV-19/USA/FL-BPHL-0680/2020\|EPI_ISL_509738\|2020- | ···················· | ···················· | ···················· | ···················· | ···················· | ···················· |
| F | 1690 | hCoV-19/USA/FL-BPHL-0681/2020\|EPI_ISL_509739\|2020- | ···················· | ···················· | ···················· | ···················· | ···················· | ···················· |
| F | 1691 | hCoV-19/USA/FL-BPHL-0682/2020\|EPI_ISL_509740\|2020- | ···················· | ···················· | ···················· | ···················· | ···················· | ···················· |
| F | 1692 | hCoV-19/USA/FL-BPHL-0683/2020\|EPI_ISL_509741\|2020- | ···················· | ···················· | ···················· | ···················· | ···················· | ···················· |
| F | 1693 | hCoV-19/USA/FL-BPHL-0684/2020\|EPI_ISL_509742\|2020- | ···················· | ···················· | ···················· | ···················· | ···················· | ···················· |
| F | 1695 | hCoV-19/USA/FL-BPHL-0687/2020\|EPI_ISL_509745\|2020- | ···················· | ···················· | ···················· | ···················· | ···················· | ···················· |
| F | 1697 | hCoV-19/USA/FL-BPHL-0689/2020\|EPI_ISL_509747\|2020- | ···················· | ···················· | ···················· | ···················· | ···················· | ···················· |
| F | 1698 | hCoV-19/USA/FL-BPHL-0690/2020\|EPI_ISL_509748\|2020- | ···················· | ···················· | ···················· | ···················· | ···················· | ···················· |
| F | 1701 | hCoV-19/USA/FL-BPHL-0693/2020\|EPI_ISL_509751\|2020- | ···················· | ···················· | ···················· | ···················· | ···················· | ···················· |
| F | 1702 | hCoV-19/USA/FL-BPHL-0694/2020\|EPI_ISL_509752\|2020- | ···················· | ···················· | ···················· | ···················· | ···················· | ···················· |
| F | 1703 | hCoV-19/USA/FL-BPHL-0695/2020\|EPI_ISL_509753\|2020- | ···················· | ···················· | ···················· | ···················· | ···················· | ···················· |
| F | 1704 | hCoV-19/USA/FL-BPHL-0696/2020\|EPI_ISL_509754\|2020- | ···················· | ···················· | ···················· | ···················· | ···················· | ···················· |
| F | 1705 | hCoV-19/USA/FL-BPHL-0697/2020\|EPI_ISL_509755\|2020- | ···················· | ···················· | ···················· | ···················· | ···················· | ···················· |
| F | 1706 | hCoV-19/USA/FL-BPHL-0698/2020\|EPI_ISL_509756\|2020- | ···················· | ···················· | ···················· | ···················· | ···················· | ···················· |
| F | 1707 | hCoV-19/USA/FL-BPHL-0699/2020\|EPI_ISL_509757\|2020- | ···················· | ···················· | ···················· | ···················· | ···················· | ···················· |
| F | 1708 | hCoV-19/USA/FL-BPHL-0700/2020\|EPI_ISL_509758\|2020- | ···················· | ···················· | ···················· | ···················· | ···················· | ···················· |
| F | 1709 | hCoV-19/USA/FL-BPHL-0701/2020\|EPI_ISL_509759\|2020- | ···················· | ···················· | ···················· | ···················· | ···················· | ···················· |
| F | 1710 | hCoV-19/South Korea/KCDC2229/2020\|EPI_ISL_510567\|2 | ···················· | ···················· | ···················· | ···················· | ···················· | ···················· |
| F | 1711 | hCoV-19/South Korea/KCDC2230/2020\|EPI_ISL_510568\|2 | ···················· | ···················· | ···················· | ···················· | ···················· | ···················· |
| F | 1712 | hCoV-19/South Korea/KCDC2231/2020\|EPI_ISL_510569\|2 | ···················· | ···················· | ···················· | ···················· | ···················· | ···················· |
| F | 1713 | hCoV-19/South Korea/KCDC2236/2020\|EPI_ISL_510574\|2 | ···················· | ···················· | ···················· | ···················· | ···················· | ···················· |
| F | 1714 | hCoV-19/South Korea/KCDC2237/2020\|EPI_ISL_510575\|2 | ···················· | ···················· | ···················· | ···················· | ···················· | ···················· |
| F | 1715 | hCoV-19/South Korea/KCDC2238/2020\|EPI_ISL_510576\|2 | ···················· | ···················· | ···················· | ···················· | ···················· | ···················· |
| F | 1716 | hCoV-19/South Korea/KCDC2240/2020\|EPI_ISL_510578\|2 | ···················· | ···················· | ···················· | ···················· | ···················· | ···················· |
| F | 1717 | hCoV-19/South Korea/KCDC2241/2020\|EPI_ISL_510579\|2 | ···················· | ···················· | ···················· | ···················· | ···················· | ···················· |
| F | 1718 | hCoV-19/South Korea/KCDC2242/2020\|EPI_ISL_510580\|2 | ···················· | ···················· | ···················· | ···················· | ···················· | ···················· |
| F | 1719 | hCoV-19/South Korea/KCDC2243/2020\|EPI_ISL_510581\|2 | ···················· | ···················· | ···················· | ···················· | ···················· | ···················· |
| F | 1720 | hCoV-19/South Korea/KCDC2244/2020\|EPI_ISL_510582\|2 | ···················· | ···················· | ···················· | ···················· | ···················· | ···················· |
| F | 1721 | hCoV-19/South Korea/KCDC2245/2020\|EPI_ISL_510583\|2 | ···················· | ···················· | ···················· | ···················· | ···················· | ···················· |
| F | 1722 | hCoV-19/South Korea/KCDC2246/2020\|EPI_ISL_510584\|2 | ···················· | ···················· | ···················· | ···················· | ···················· | ···················· |
| F | 1723 | hCoV-19/South Korea/KCDC2247/2020\|EPI_ISL_510585\|2 | ···················· | ···················· | ···················· | ···················· | ···················· | ···················· |
| F | 1724 | hCoV-19/South Korea/KCDC2248/2020\|EPI_ISL_510586\|2 | ···················· | ···················· | ···················· | ···················· | ···················· | ···················· |
| F | 1725 | hCoV-19/South Korea/KCDC2249/2020\|EPI_ISL_510587\|2 | ···················· | ···················· | ···················· | ···················· | ···················· | ···················· |
| F | 1726 | hCoV-19/South Korea/KCDC2250/2020\|EPI_ISL_510588\|2 | ···················· | ···················· | ···················· | ···················· | ···················· | ···················· |
| F | 1727 | hCoV-19/South Korea/KCDC2251/2020\|EPI_ISL_510589\|2 | ···················· | ···················· | ···················· | ···················· | ···················· | ···················· |
| F | 1728 | hCoV-19/South Korea/KCDC2252/2020\|EPI_ISL_510590\|2 | ···················· | ···················· | ···················· | ···················· | ···················· | ···················· |
| F | 1729 | hCoV-19/South Korea/KCDC2253/2020\|EPI_ISL_510591\|2 | ···················· | ···················· | ···················· | ···················· | ···················· | ···················· |
| F | 1730 | hCoV-19/South Korea/KCDC2254/2020\|EPI_ISL_510592\|2 | ···················· | ···················· | ···················· | ···················· | ···················· | ···················· |
| F | 1731 | hCoV-19/South Korea/KCDC2255/2020\|EPI_ISL_510593\|2 | ···················· | ···················· | ···················· | ···················· | ···················· | ···················· |
| F | 1732 | hCoV-19/South Korea/KCDC2256/2020\|EPI_ISL_510594\|2 | ···················· | ···················· | ···················· | ···················· | ···················· | ···················· |
| F | 1733 | hCoV-19/South Korea/KCDC2257/2020\|EPI_ISL_510595\|2 | ···················· | ···················· | ···················· | ···················· | ···················· | ···················· |
| F | 1734 | hCoV-19/South Korea/KCDC2258/2020\|EPI_ISL_510596\|2 | ···················· | ···················· | ···················· | ···················· | ···················· | ···················· |
| F | 1735 | hCoV-19/South Korea/KCDC2259/2020\|EPI_ISL_510597\|2 | ···················· | ···················· | ···················· | ···················· | ···················· | ···················· |
| F | 1736 | hCoV-19/South Korea/KCDC2260/2020\|EPI_ISL_510598\|2 | ···················· | ···················· | ···················· | ···················· | ···················· | ···················· |
| F | 1737 | hCoV-19/South Korea/KCDC2261/2020\|EPI_ISL_510599\|2 | ···················· | ···················· | ···················· | ···················· | ···················· | ···················· |
| F | 1738 | hCoV-19/Switzerland/BL-ETHZ-200012/2020\|EPI_ISL_51 | ···················· | ···················· | ···················· | ···················· | ···················· | ···················· |
| F | 1739 | hCoV-19/Switzerland/UR-ETHZ-200013/2020\|EPI_ISL_51 | ···················· | ·A·················· | ···················· | ···················· | ···················· | ···················· |
| F | 1740 | hCoV-19/Switzerland/UR-ETHZ-200014/2020\|EPI_ISL_51 | ···················· | ·A·················· | ···················· | ···················· | ···················· | ···················· |
| F | 1741 | hCoV-19/Switzerland/BL-ETHZ-200015/2020\|EPI_ISL_51 | ···················· | ···················· | ···················· | ···················· | ···················· | ···················· |
| F | 1742 | hCoV-19/Switzerland/ZH-ETHZ-200016/2020\|EPI_ISL_51 | ···················· | ···················· | ···················· | ···················· | ···················· | ···················· |
| F | 1743 | hCoV-19/Switzerland/ZH-ETHZ-200017/2020\|EPI_ISL_51 | ···················· | ···················· | ···················· | ···················· | ···················· | ···················· |
| F | 1744 | hCoV-19/Switzerland/BL-ETHZ-200018/2020\|EPI_ISL_51 | ···················· | ···················· | ···················· | ···················· | ···················· | ···················· |
| F | 1745 | hCoV-19/Switzerland/ZH-ETHZ-200019/2020\|EPI_ISL_51 | ···················· | ···················· | ···················· | ···················· | ···················· | ···················· |
| F | 1746 | hCoV-19/Switzerland/BE-ETHZ-200020/2020\|EPI_ISL_51 | ···················· | ···················· | ···················· | ···················· | ···················· | ···················· |
| F | 1747 | hCoV-19/Switzerland/BE-ETHZ-200021/2020\|EPI_ISL_51 | ···················· | ···················· | ···················· | ···················· | ···················· | ···················· |
| F | 1748 | hCoV-19/Switzerland/TI-ETHZ-200022/2020\|EPI_ISL_51 | ···················· | ···················· | ···················· | ···················· | ···················· | ···················· |
| F | 1749 | hCoV-19/Switzerland/AG-ETHZ-200023/2020\|EPI_ISL_51 | ···················· | ···················· | ···················· | ···················· | ···················· | ···················· |
| F | 1750 | hCoV-19/Switzerland/BL-ETHZ-200024/2020\|EPI_ISL_51 | ···················· | ···················· | ···················· | ···················· | ···················· | ···················· |
| F | 1751 | hCoV-19/Switzerland/BE-ETHZ-200025/2020\|EPI_ISL_51 | ···················· | ···················· | ···················· | ···················· | ···················· | ···················· |
| F | 1752 | hCoV-19/Switzerland/ZH-ETHZ-200026/2020\|EPI_ISL_51 | ···················· | ···················· | ···················· | ···················· | ···················· | ···················· |
| F | 1753 | hCoV-19/Switzerland/ZH-ETHZ-200027/2020\|EPI_ISL_51 | ···················· | ···················· | ···················· | ···················· | ···················· | ···················· |
| F | 1754 | hCoV-19/Switzerland/ZH-ETHZ-200028/2020\|EPI_ISL_51 | ···················· | ···················· | ···················· | ···················· | ···················· | ···················· |
| F | 1755 | hCoV-19/Switzerland/SG-ETHZ-200029/2020\|EPI_ISL_51 | ···················· | ···················· | ···················· | ···················· | ···················· | ···················· |
| F | 1756 | hCoV-19/Switzerland/AG-ETHZ-200030/2020\|EPI_ISL_51 | ···················· | ···················· | ···················· | ···················· | ···················· | ···················· |
| F | 1757 | hCoV-19/Switzerland/VD-ETHZ-200031/2020\|EPI_ISL_51 | ···················· | ···················· | ···················· | ···················· | ···················· | ···················· |
| F | 1758 | hCoV-19/Switzerland/BE-ETHZ-200032/2020\|EPI_ISL_51 | ···················· | ···················· | ···················· | ···················· | ···················· | ···················· |
| F | 1759 | hCoV-19/Switzerland/ZH-ETHZ-200033/2020\|EPI_ISL_51 | ···················· | ···················· | ···················· | ···················· | ···················· | ···················· |
| F | 1760 | hCoV-19/Switzerland/BE-ETHZ-200035/2020\|EPI_ISL_51 | ···················· | ···················· | ···················· | ···················· | ···················· | ···················· |
| F | 1761 | hCoV-19/Switzerland/BL-ETHZ-200036/2020\|EPI_ISL_51 | ···················· | ···················· | ···················· | ···················· | ···················· | ···················· |
| F | 1762 | hCoV-19/Switzerland/ZH-ETHZ-200037/2020\|EPI_ISL_51 | ···················· | ···················· | ···················· | ···················· | ···················· | ···················· |
| F | 1763 | hCoV-19/Switzerland/BL-ETHZ-200038/2020\|EPI_ISL_51 | ···················· | ···················· | ···················· | ···················· | ···················· | ···················· |
| F | 1764 | hCoV-19/Switzerland/BL-ETHZ-200039/2020\|EPI_ISL_51 | ···················· | ···················· | ···················· | ···················· | ···················· | ···················· |

```
F   1765 hCoV-19/Switzerland/GE-ETHZ-200042/2020|EPI_ISL_51   ····················  ····················  ·····················  ····················  ····················  ····················
F   1766 hCoV-19/Switzerland/BE-ETHZ-200043/2020|EPI_ISL_51   ····················  ····················  ·····················  ····················  ····················  ····················
F   1767 hCoV-19/Switzerland/BL-ETHZ-200045/2020|EPI_ISL_51   ···········T········  ····················  ·····················  ····················  ····················  ····················
F   1768 hCoV-19/Switzerland/SZ-ETHZ-200046/2020|EPI_ISL_51   ····················  ····················  ·····················  ····················  ····················  ····················
F   1769 hCoV-19/Switzerland/ZH-ETHZ-200047/2020|EPI_ISL_51   ····················  ····················  ·····················  ····················  ····················  ····················
F   1770 hCoV-19/Switzerland/ZH-ETHZ-200048/2020|EPI_ISL_51   ····················  ····················  ·····················  ····················  ····················  ····················
F   1771 hCoV-19/Switzerland/SZ-ETHZ-200049/2020|EPI_ISL_51   ····················  ····················  ·····················  ····················  ····················  ····················
F   1772 hCoV-19/Switzerland/SH-ETHZ-200050/2020|EPI_ISL_51   ····················  ····················  ·····················  ····················  ····················  ····················
F   1773 hCoV-19/Switzerland/BL-ETHZ-200052/2020|EPI_ISL_51   ····················  ····················  ·····················  ····················  ····················  ····················
F   1774 hCoV-19/Switzerland/BL-ETHZ-200053/2020|EPI_ISL_51   ···········T········  ····················  ·····················  ····················  ····················  ····················
F   1775 hCoV-19/Switzerland/ZH-ETHZ-200054/2020|EPI_ISL_51   ····················  ····················  ·····················  ····················  ····················  ····················
F   1776 hCoV-19/Switzerland/BS-ETHZ-200055/2020|EPI_ISL_51   ····················  ····················  ·····················  ····················  ····················  ····················
F   1777 hCoV-19/Switzerland/BL-ETHZ-200056/2020|EPI_ISL_51   ····················  ····················  ·····················  ····················  ····················  ····················
F   1778 hCoV-19/Switzerland/BL-ETHZ-200059/2020|EPI_ISL_51   ····················  ····················  ·····················  ····················  ····················  ····················
F   1779 hCoV-19/Switzerland/BE-ETHZ-200060/2020|EPI_ISL_51   ····················  ·A··················  ·····················  ····················  ····················  ····················
F   1780 hCoV-19/Switzerland/VD-ETHZ-200069/2020|EPI_ISL_51   ····················  ····················  ·····················  ····················  ····················  ····················
F   1781 hCoV-19/Switzerland/VD-ETHZ-200070/2020|EPI_ISL_51   ····················  ····················  ·····················  ····················  ····················  ····················
F   1782 hCoV-19/Switzerland/ZH-ETHZ-210004/2020|EPI_ISL_51   ····················  ····················  ·····················  ····················  ······K·············  ····················
F   1783 hCoV-19/Vietnam/VNHN_5152/2020|EPI_ISL_511898|2020   ····················  ····················  ·····················  ····················  ····················  ····················
F   1784 hCoV-19/India/WB-S83/2020|EPI_ISL_511899|2020-05-2   ····················  ····················  ·····················  ····················  ····················  ····················
F   1786 hCoV-19/Israel/CVL-n26150/2020|EPI_ISL_514291|2020   ····················  ·A··················  ·····················  ····················  ····················  ····················
F   1787 hCoV-19/Israel/CVL-n26156/2020|EPI_ISL_514292|2020   ····················  ·A··················  ·····················  ····················  ····················  ····················
F   1788 hCoV-19/Israel/CVL-n26166/2020|EPI_ISL_514293|2020   ····················  ····················  ·····················  ····················  ····················  ····················
F   1789 hCoV-19/Israel/CVL-n26167/2020|EPI_ISL_514294|2020   ····················  ····················  ···········T·········  ····················  ····················  ····················
F   1790 hCoV-19/Israel/CVL-n26168/2020|EPI_ISL_514295|2020   ····················  ·A··················  ·····················  ····················  ····················  ····················
F   1792 hCoV-19/Israel/CVL-n26412/2020|EPI_ISL_514299|2020   ····················  ····················  ···········T·········  ····················  ····················  ····················
F   1793 hCoV-19/USA/WA-S2368/2020|EPI_ISL_497000|2020-06-1   ····················  ····················  ·····················  ····················  ····················  ····················
F   1794 hCoV-19/USA/WA-S2371/2020|EPI_ISL_497003|2020-06-1   ····················  ····················  ·····················  ····················  ····················  ····················
F   1795 hCoV-19/USA/WA-S2372/2020|EPI_ISL_497004|2020-06-1   ····················  ····················  ·····················  ····················  ····················  ····················
F   1796 hCoV-19/USA/WA-S2373/2020|EPI_ISL_497005|2020-06-1   ····················  ····················  ·····················  ····················  ····················  ····················
F   1797 hCoV-19/USA/WA-S2374/2020|EPI_ISL_497006|2020-06-1   ····················  ····················  ·····················  ····················  ····················  ····················
F   1798 hCoV-19/USA/WA-S2375/2020|EPI_ISL_497007|2020-06-1   ····················  ····················  ·····················  ····················  ····················  ····················
F   1799 hCoV-19/USA/WA-S2376/2020|EPI_ISL_497008|2020-06-1   ····················  ····················  ·····················  ····················  ····················  ····················
F   1800 hCoV-19/USA/WA-S2377/2020|EPI_ISL_497009|2020-06-1   ····················  ····················  ·····················  ····················  ····················  ····················
F   1801 hCoV-19/USA/WA-S2378/2020|EPI_ISL_497010|2020-06-1   ····················  ····················  ·····················  ····················  ····················  ····················
F   1802 hCoV-19/USA/WA-S2379/2020|EPI_ISL_497011|2020-06-1   ····················  ····················  ·····················  ····················  ····················  ····················
F   1803 hCoV-19/USA/WA-S2380/2020|EPI_ISL_497012|2020-06-1   ····················  ····················  ·····················  ····················  ····················  ····················
F   1804 hCoV-19/USA/WA-S2381/2020|EPI_ISL_497013|2020-06-1   ····················  ····················  ·····················  ····················  ····················  ····················
F   1805 hCoV-19/USA/WA-S2382/2020|EPI_ISL_497014|2020-06-1   ····················  ····················  ·····················  ····················  ····················  ····················
F   1806 hCoV-19/USA/WA-S2383/2020|EPI_ISL_497015|2020-06-1   ····················  ····················  ·····················  ····················  ····················  ····················
F   1807 hCoV-19/USA/WA-S2384/2020|EPI_ISL_497016|2020-06-1   ····················  ····················  ·····················  ····················  ····················  ····················
F   1808 hCoV-19/USA/WA-S2385/2020|EPI_ISL_497017|2020-06-1   ····················  ····················  ·····················  ····················  ····················  ····················
F   1809 hCoV-19/USA/WA-S2386/2020|EPI_ISL_497018|2020-06-1   ····················  ····················  ·····················  ····················  ····················  ····················
F   1810 hCoV-19/USA/WA-S2387/2020|EPI_ISL_497019|2020-06-1   ····················  ····················  ·····················  ····················  ····················  ····················
F   1811 hCoV-19/USA/WA-S2388/2020|EPI_ISL_497020|2020-06-1   ····················  ····A···············  ·····················  ····················  ····················  ····················
F   1812 hCoV-19/USA/WA-S2389/2020|EPI_ISL_497021|2020-06-1   ····················  ····················  ·····················  ····················  ····················  ····················
F   1813 hCoV-19/USA/WA-S2390/2020|EPI_ISL_497022|2020-06-1   ····················  ····················  ·····················  ····················  ····················  ····················
F   1814 hCoV-19/USA/WA-S2391/2020|EPI_ISL_497023|2020-06-1   ····················  ····················  ·····················  ····················  ····················  ····················
F   1815 hCoV-19/USA/WA-S2392/2020|EPI_ISL_497024|2020-06-1   ····················  ····················  ·····················  ····················  ····················  ····················
F   1816 hCoV-19/USA/WA-S2393/2020|EPI_ISL_497025|2020-06-1   ····················  ····················  ·····················  ····················  ····················  ····················
F   1817 hCoV-19/USA/WA-S2394/2020|EPI_ISL_497026|2020-06-1   ····················  ····················  ·····················  ····················  ····················  ····················
F   1818 hCoV-19/USA/WA-S2395/2020|EPI_ISL_497027|2020-06-1   ····················  ····················  ·····················  ····················  ····················  ····················
F   1819 hCoV-19/USA/WA-S2396/2020|EPI_ISL_497028|2020-06-1   ····················  ····················  ·····················  ····················  ····················  ····················
F   1820 hCoV-19/USA/WA-S2397/2020|EPI_ISL_497029|2020-06-1   ····················  ····················  ·····················  ····················  ····················  ····················
F   1821 hCoV-19/USA/WA-S2398/2020|EPI_ISL_497030|2020-06-1   ····················  ····················  ·····················  ····················  ····················  ····················
F   1822 hCoV-19/USA/WA-S2399/2020|EPI_ISL_497031|2020-06-1   ····················  ····················  ·····················  ····················  ····················  ····················
F   1823 hCoV-19/USA/WA-S2400/2020|EPI_ISL_497032|2020-06-1   ····················  ····················  ·····················  ····················  ····················  ····················
F   1824 hCoV-19/USA/WA-S2401/2020|EPI_ISL_497033|2020-06-1   ····················  ····················  ·····················  ····················  ····················  ····················
F   1825 hCoV-19/USA/WA-S2402/2020|EPI_ISL_497034|2020-06-1   ····················  ····················  ·····················  ····················  ····················  ····················
F   1826 hCoV-19/USA/WA-S2403/2020|EPI_ISL_497035|2020-06-1   ····················  ····················  ·····················  ····················  ····················  ····················
F   1827 hCoV-19/USA/WA-S2404/2020|EPI_ISL_497036|2020-06-1   ····················  ····················  ·····················  ····················  ····················  ····················
F   1828 hCoV-19/USA/WA-S2405/2020|EPI_ISL_497037|2020-06-1   ····················  ····················  ·····················  ····················  ····················  ····················
F   1829 hCoV-19/USA/WA-S2406/2020|EPI_ISL_497038|2020-06-1   ····················  ····················  ·····················  ····················  ····················  ····················
F   1830 hCoV-19/USA/WA-S2407/2020|EPI_ISL_497039|2020-06-1   ····················  ····················  ·····················  ····················  ····················  ····················
F   1831 hCoV-19/USA/WA-S2408/2020|EPI_ISL_497040|2020-06-1   ····················  ····A···············  ·····················  ····················  ····················  ····················
F   1832 hCoV-19/USA/WA-S2409/2020|EPI_ISL_497041|2020-06-1   ····················  ····················  ·····················  ····················  ····················  ····················
F   1833 hCoV-19/USA/WA-S2410/2020|EPI_ISL_497042|2020-06-1   ····················  ····················  ·····················  ····················  ····················  ····················
F   1834 hCoV-19/USA/WA-S2411/2020|EPI_ISL_497043|2020-06-1   ····················  ····················  ·····················  ····················  ····················  ····················
F   1835 hCoV-19/USA/WA-S2412/2020|EPI_ISL_497044|2020-06-1   ····················  ····················  ·····················  ····················  ····················  ····················
F   1836 hCoV-19/USA/WA-S2413/2020|EPI_ISL_497045|2020-06-1   ····················  ····················  ·····················  ····················  ····················  ····················
F   1837 hCoV-19/USA/WA-S2414/2020|EPI_ISL_497046|2020-06-1   ····················  ····················  ·····················  ····················  ····················  ····················
F   1838 hCoV-19/USA/WA-S2445/2020|EPI_ISL_497077|2020-06-1   ····················  ····················  ·····················  ····················  ····················  ····················
F   1839 hCoV-19/USA/WA-S2446/2020|EPI_ISL_497078|2020-06-1   ····················  ····················  ·····················  ····················  ····················  ····················
F   1840 hCoV-19/USA/WA-S2447/2020|EPI_ISL_497079|2020-06-1   ····················  ····················  ·····················  ····················  ····················  ····················
F   1841 hCoV-19/England/LIVE-A5841/2020|EPI_ISL_499640|202   ····················  ····················  ·····················  ····················  ····················  ····················
F   1842 hCoV-19/England/LIVE-A587E/2020|EPI_ISL_499641|202   ····················  ····················  ·····················  ····················  ····················  ····················
F   1843 hCoV-19/England/LIVE-A58BA/2020|EPI_ISL_499642|202   ····················  ····················  ·····················  ····················  ····················  ····················
F   1844 hCoV-19/England/LIVE-A58C9/2020|EPI_ISL_499643|202   ····················  ····················  ·····················  ····················  ····················  ····················
F   1845 hCoV-19/England/LIVE-A5920/2020|EPI_ISL_499648|202   ····················  ····················  ·····················  ····················  ····················  ····················
F   1846 hCoV-19/England/LIVE-A593F/2020|EPI_ISL_499649|202   ····················  ····················  ·····················  ····················  ····················  ····················
```

| | | | | | | | |
|---|---|---|---|---|---|---|---|
| F | 1847 hCoV-19/England/LIVE-A5A96/2020\|EPI_ISL_499655\|202 | .................... | .................... | .................... | .................... | .................... | .................... |
| F | 1851 hCoV-19/England/LIVE-A5D24/2020\|EPI_ISL_499659\|202 | .................... | .................... | .................... | .................... | .................... | .................... |
| F | 1852 hCoV-19/England/LIVE-A5D33/2020\|EPI_ISL_499660\|202 | .................... | .................... | .................... | .................... | .................... | .................... |
| F | 1853 hCoV-19/England/LIVE-A5D7F/2020\|EPI_ISL_499661\|202 | .................... | .................... | .................... | .................... | .................... | .................... |
| F | 1856 hCoV-19/England/LIVE-A5EB8/2020\|EPI_ISL_499664\|202 | .................... | .................... | .................... | .................... | .................... | .................... |
| F | 1857 hCoV-19/England/LIVE-A5ED6/2020\|EPI_ISL_499665\|202 | .................... | .................... | .................... | .................... | .................... | .................... |
| F | 1858 hCoV-19/England/LIVE-A5EE5/2020\|EPI_ISL_499666\|202 | .................... | .................... | .................... | .................... | .................... | .................... |
| F | 1859 hCoV-19/England/LIVE-A5F00/2020\|EPI_ISL_499667\|202 | .................... | .................... | .................... | .................... | .................... | .................... |
| F | 1860 hCoV-19/England/LIVE-A5F1F/2020\|EPI_ISL_499668\|202 | .................... | .................... | .................... | .................... | .................... | .................... |
| F | 1861 hCoV-19/England/LIVE-A5F97/2020\|EPI_ISL_499669\|202 | .................... | .................... | .................... | .................... | .................... | .................... |
| F | 1862 hCoV-19/England/LIVE-A5FA6/2020\|EPI_ISL_499670\|202 | .................... | .................... | .................... | .................... | .................... | .................... |
| F | 1863 hCoV-19/England/LIVE-A5FC4/2020\|EPI_ISL_499671\|202 | .................... | .................... | .................... | .................... | .................... | .................... |
| F | 1864 hCoV-19/England/LIVE-A5FD3/2020\|EPI_ISL_499672\|202 | .................... | .................... | .................... | .................... | .................... | .................... |
| F | 1865 hCoV-19/England/LIVE-A5FE2/2020\|EPI_ISL_499673\|202 | .................... | .................... | .................... | .................... | .................... | .................... |
| F | 1866 hCoV-19/England/LIVE-A600C/2020\|EPI_ISL_499674\|202 | .................... | .................... | .................... | .................... | .................... | .................... |
| F | 1867 hCoV-19/England/LIVE-A601B/2020\|EPI_ISL_499675\|202 | .................... | .................... | .................... | .................... | .................... | .................... |
| F | 1868 hCoV-19/England/LIVE-A602A/2020\|EPI_ISL_499676\|202 | .................... | .................... | .................... | .................... | .................... | .................... |
| F | 1869 hCoV-19/England/LIVE-A6039/2020\|EPI_ISL_499677\|202 | .................... | .................... | .................... | .................... | .................... | .................... |
| F | 1870 hCoV-19/England/LIVE-A6048/2020\|EPI_ISL_499678\|202 | .................... | .................... | .................... | .................... | .................... | .................... |
| F | 1871 hCoV-19/England/LIVE-A6066/2020\|EPI_ISL_499679\|202 | .................... | .................... | .................... | .................... | .................... | .................... |
| F | 1872 hCoV-19/England/LIVE-A6075/2020\|EPI_ISL_499680\|202 | .................... | .................... | .................... | .................... | .................... | .................... |
| F | 1873 hCoV-19/England/LIVE-A6093/2020\|EPI_ISL_499681\|202 | .................... | .................... | .................... | .................... | .................... | .................... |
| F | 1874 hCoV-19/England/LIVE-A60B1/2020\|EPI_ISL_499682\|202 | .................... | .................... | .................... | .................... | .................... | .................... |
| F | 1876 hCoV-19/England/LIVE-A60EE/2020\|EPI_ISL_499684\|202 | .................... | .................... | .................... | .................... | .................... | .................... |
| F | 1877 hCoV-19/England/LIVE-A60FD/2020\|EPI_ISL_499685\|202 | .................... | .................... | .................... | .................... | .................... | .................... |
| F | 1878 hCoV-19/England/LIVE-A6109/2020\|EPI_ISL_499686\|202 | .................... | .................... | .................... | .................... | .................... | .................... |
| F | 1879 hCoV-19/England/LIVE-A6163/2020\|EPI_ISL_499687\|202 | .................... | .................... | .................... | .................... | .................... | .................... |
| F | 1881 hCoV-19/England/LIVE-A6181/2020\|EPI_ISL_499689\|202 | .................... | .................... | .................... | .................... | .................... | .................... |
| F | 1882 hCoV-19/England/LIVE-A6190/2020\|EPI_ISL_499690\|202 | .................... | .................... | .................... | .................... | .................... | .................... |
| F | 1883 hCoV-19/England/LIVE-A61CD/2020\|EPI_ISL_499691\|202 | .................... | .................... | .................... | .................... | .................... | .................... |
| F | 1884 hCoV-19/England/LIVE-A61DC/2020\|EPI_ISL_499692\|202 | .................... | .................... | .................... | .................... | .................... | .................... |
| F | 1885 hCoV-19/England/LIVE-A61EB/2020\|EPI_ISL_499693\|202 | .................... | .................... | .................... | .................... | .................... | .................... |
| F | 1886 hCoV-19/England/LIVE-A6215/2020\|EPI_ISL_499694\|202 | .................... | .................... | .................... | .................... | .................... | .................... |
| F | 1887 hCoV-19/England/LIVE-A6233/2020\|EPI_ISL_499695\|202 | .................... | .A.................. | .................... | .................... | .................... | .................... |
| F | 1888 hCoV-19/England/LIVE-A6242/2020\|EPI_ISL_499696\|202 | .................... | .................... | .................... | .................... | .................... | .................... |
| F | 1889 hCoV-19/England/LIVE-A6260/2020\|EPI_ISL_499697\|202 | .................... | .................... | .................... | .................... | .................... | .................... |
| F | 1890 hCoV-19/England/LIVE-A628E/2020\|EPI_ISL_499698\|202 | .................... | .................... | .................... | .................... | .................... | .................... |
| F | 1891 hCoV-19/England/LIVE-A62AC/2020\|EPI_ISL_499699\|202 | .................... | .................... | .................... | .................... | .................... | .................... |
| F | 1892 hCoV-19/India/MH-GA14/2020\|EPI_ISL_508210\|2020-06- | .................... | .................... | .................... | .................... | .................... | .................... |
| F | 1893 hCoV-19/India/MH-GA15/2020\|EPI_ISL_508211\|2020-06- | .................... | .................... | .................... | .................... | .................... | .................... |
| F | 1894 hCoV-19/India/MH-GA16/2020\|EPI_ISL_508212\|2020-06- | .................... | .................... | .................... | .................... | .................... | .................... |
| F | 1895 hCoV-19/India/MH-GA17/2020\|EPI_ISL_508213\|2020-06- | .................... | .................... | .................... | .................... | .................... | .................... |
| F | 1896 hCoV-19/India/MH-GA19/2020\|EPI_ISL_508214\|2020-06- | .................... | .................... | .................... | .................... | .................... | .................... |
| F | 1897 hCoV-19/India/MH-GA2/2020\|EPI_ISL_508215\|2020-06-1 | .................... | .................... | .................... | .................... | .................... | .................... |
| F | 1898 hCoV-19/India/MH-GA20/2020\|EPI_ISL_508216\|2020-06- | .................... | .................... | .................... | .................... | .................... | .................... |
| F | 1899 hCoV-19/India/MH-GA21/2020\|EPI_ISL_508217\|2020-06- | .................... | .................... | .................... | .................... | .................... | .................... |
| F | 1900 hCoV-19/India/MH-GA22/2020\|EPI_ISL_508218\|2020-06- | .................... | .................... | .................... | .................... | .................... | .................... |
| F | 1901 hCoV-19/India/MH-GA24/2020\|EPI_ISL_508219\|2020-06- | .................... | .................... | .................... | .................... | .................... | .................... |
| F | 1902 hCoV-19/India/MH-GA25/2020\|EPI_ISL_508220\|2020-06- | .................... | .................... | .................... | .................... | .................... | .................... |
| F | 1903 hCoV-19/India/MH-GA26/2020\|EPI_ISL_508221\|2020-06- | .................... | .................... | .................... | .................... | .................... | .................... |
| F | 1904 hCoV-19/India/MH-GA27/2020\|EPI_ISL_508222\|2020-06- | .................... | .................... | .................... | .................... | .................... | .................... |
| F | 1905 hCoV-19/India/MH-GA28/2020\|EPI_ISL_508223\|2020-06- | .................... | .................... | .................... | .................... | .................... | .................... |
| F | 1906 hCoV-19/India/MH-GA29/2020\|EPI_ISL_508224\|2020-06- | .................... | .................... | .................... | .................... | .................... | .................... |
| F | 1907 hCoV-19/India/MH-GA31/2020\|EPI_ISL_508225\|2020-06- | .................... | .................... | .................... | .................... | .................... | .................... |
| F | 1908 hCoV-19/India/MH-GA32/2020\|EPI_ISL_508226\|2020-06- | .................... | .................... | .................... | .................... | .................... | .................... |
| F | 1909 hCoV-19/India/MH-GA33/2020\|EPI_ISL_508227\|2020-06- | .................... | .................... | .................... | .................... | .................... | .................... |
| F | 1910 hCoV-19/India/MH-GA34/2020\|EPI_ISL_508228\|2020-06- | .................... | .................... | .................... | .................... | .................... | .................... |
| F | 1911 hCoV-19/India/MH-GA35/2020\|EPI_ISL_508229\|2020-06- | .................... | .................... | .................... | .................... | .................... | .................... |
| F | 1912 hCoV-19/India/MH-GA36/2020\|EPI_ISL_508230\|2020-06- | .................... | .................... | .................... | .................... | .................... | .................... |
| F | 1913 hCoV-19/India/MH-GA37/2020\|EPI_ISL_508231\|2020-06- | .................... | .................... | .................... | .................... | .................... | .................... |
| F | 1914 hCoV-19/India/MH-GA38/2020\|EPI_ISL_508232\|2020-06- | .................... | .................... | .................... | .................... | .................... | .................... |
| F | 1915 hCoV-19/India/MH-GA39/2020\|EPI_ISL_508233\|2020-06- | .................... | .................... | .................... | .................... | .................... | .................... |
| F | 1916 hCoV-19/India/MH-GA4/2020\|EPI_ISL_508234\|2020-06-1 | .................... | .................... | .................... | .................... | .................... | .................... |
| F | 1917 hCoV-19/India/MH-GA40/2020\|EPI_ISL_508235\|2020-06- | .................... | .................... | .................... | .................... | .................... | .................... |
| F | 1918 hCoV-19/India/MH-GA41/2020\|EPI_ISL_508236\|2020-06- | .................... | .................... | .................... | .................... | .................... | .................... |
| F | 1919 hCoV-19/India/MH-GA42/2020\|EPI_ISL_508237\|2020-06- | .................... | .................... | .................... | .................... | .................... | .................... |
| F | 1920 hCoV-19/India/MH-GA43/2020\|EPI_ISL_508238\|2020-06- | .................... | .................... | .................... | .................... | .................... | .................... |
| F | 1921 hCoV-19/India/MH-GA45/2020\|EPI_ISL_508239\|2020-06- | .................... | .................... | .................... | .................... | .................... | .................... |
| F | 1922 hCoV-19/India/MH-GA46/2020\|EPI_ISL_508240\|2020-06- | .................... | .................... | .................... | .................... | .................... | .................... |
| F | 1923 hCoV-19/India/MH-GA47/2020\|EPI_ISL_508241\|2020-06- | .................... | .................... | .................... | .................... | .................... | .................... |
| F | 1924 hCoV-19/India/MH-GA48/2020\|EPI_ISL_508242\|2020-06- | .................... | .................... | .................... | .................... | .................... | .................... |
| F | 1925 hCoV-19/India/MH-GA49/2020\|EPI_ISL_508243\|2020-06- | .................... | .................... | .................... | .................... | .................... | .................... |
| F | 1926 hCoV-19/India/MH-GA5/2020\|EPI_ISL_508244\|2020-06-1 | .................... | .................... | .................... | .................... | .................... | .................... |
| F | 1927 hCoV-19/India/MH-GA50/2020\|EPI_ISL_508245\|2020-06- | .................... | .................... | .................... | .................... | .................... | .................... |
| F | 1928 hCoV-19/India/MH-GA51/2020\|EPI_ISL_508246\|2020-06- | .................... | .................... | .................... | .................... | .................... | .................... |
| F | 1929 hCoV-19/India/MH-GA52/2020\|EPI_ISL_508247\|2020-06- | .................... | .................... | .................... | .................... | .................... | .................... |
| F | 1930 hCoV-19/India/MH-GA53/2020\|EPI_ISL_508248\|2020-06- | .................... | .................... | .................... | .................... | .................... | .................... |
| F | 1931 hCoV-19/India/MH-GA54/2020\|EPI_ISL_508249\|2020-06- | .................... | .................... | .................... | .................... | .................... | .................... |
| F | 1932 hCoV-19/USA/UT-02915/2020\|EPI_ISL_509540\|2020-04-3 | .................... | .................... | .................... | .................... | .................... | .................... |
| F | 1933 hCoV-19/USA/UT-02918/2020\|EPI_ISL_509541\|2020-04-3 | .................... | .................... | .................... | .................... | .................... | .................... |

| | | | | | | | | |
|---|---|---|---|---|---|---|---|---|
| F | 1934 | hCoV-19/USA/UT-02928/2020\|EPI_ISL_509547\|2020-04-3 | .................... | .................... | .................... | .................... | .................... | .................... |
| F | 1935 | hCoV-19/USA/UT-02937/2020\|EPI_ISL_509549\|2020-04-3 | .................... | .................... | .................... | .................... | .................... | .................... |
| F | 1936 | hCoV-19/USA/UT-02946/2020\|EPI_ISL_509550\|2020-04-3 | .................... | .................... | .................... | .................... | .................... | .................... |
| F | 1937 | hCoV-19/USA/UT-02960/2020\|EPI_ISL_509553\|2020-05-0 | .................... | .................... | .................... | .................... | .................... | .................... |
| F | 1938 | hCoV-19/USA/UT-02962/2020\|EPI_ISL_509554\|2020-05-0 | .................... | .................... | .................... | .................... | .................... | .................... |
| F | 1939 | hCoV-19/USA/UT-03008/2020\|EPI_ISL_509558\|2020-04-3 | .................... | .................... | .................... | .................... | .................... | .................... |
| F | 1940 | hCoV-19/USA/UT-03017/2020\|EPI_ISL_509559\|2020-05-0 | .................... | .................... | .................... | .................... | .................... | .................... |
| F | 1941 | hCoV-19/USA/UT-03018/2020\|EPI_ISL_509560\|2020-05-0 | .................... | .................... | .................... | .................... | .................... | .................... |
| F | 1942 | hCoV-19/USA/UT-03021/2020\|EPI_ISL_509563\|2020-05-0 | .................... | .................... | .................... | .................... | .................... | .................... |
| F | 1943 | hCoV-19/USA/UT-01817/2020\|EPI_ISL_509568\|2020-04-1 | .................... | .................... | .................... | .................... | .................... | .................... |
| F | 1944 | hCoV-19/USA/UT-01820/2020\|EPI_ISL_509571\|2020-04-1 | .................... | .................... | .................... | .................... | .................... | .................... |
| F | 1945 | hCoV-19/USA/UT-01822/2020\|EPI_ISL_509572\|2020-04-1 | .................... | .................... | .................... | .................... | .................... | .................... |
| F | 1946 | hCoV-19/USA/UT-03447/2020\|EPI_ISL_509576\|2020-04-2 | .................... | .................... | .................... | .................... | .................... | .................... |
| F | 1947 | hCoV-19/USA/UT-03452/2020\|EPI_ISL_509577\|2020-04-1 | .................... | .................... | .................... | .................... | .................... | .................... |
| F | 1949 | hCoV-19/Egypt/HCoV2-Egy-002/2020\|EPI_ISL_510532\|20 | .................... | .................... | .................... | .................... | .................... | .................... |
| F | 1950 | hCoV-19/Brazil/PR-LRV-01/2020\|EPI_ISL_510535\|2020- | .................... | .................... | .................... | .................... | .................... | .................... |
| F | 1951 | hCoV-19/Brazil/RJ-DCVN5/2020\|EPI_ISL_510536\|2020-0 | .................... | .................... | .................... | .................... | .................... | .................... |
| F | 1952 | hCoV-19/Hong Kong/HKU-200723-107/2020\|EPI_ISL_5105 | .................... | .................... | .................... | .................... | .................... | .................... |
| F | 1953 | hCoV-19/Hong Kong/HKU-200723-108/2020\|EPI_ISL_5105 | .................... | .................... | .................... | .................... | .................... | .................... |
| F | 1954 | hCoV-19/Brazil/RJ-DCVN4/2020\|EPI_ISL_510541\|2020-0 | .................... | .................... | .................... | .................... | .................... | .................... |
| F | 1955 | hCoV-19/Australia/SAP476/2020\|EPI_ISL_510542\|2020- | .................... | .................... | .................... | .................... | .................... | .................... |
| F | 1956 | hCoV-19/Australia/SAP480/2020\|EPI_ISL_510544\|2020- | .................... | .................... | .................... | .................... | .................... | .................... |
| F | 1957 | hCoV-19/South Korea/KCDC2203/2020\|EPI_ISL_510547\|2 | .................... | .................... | .................... | .................... | .................... | .................... |
| F | 1958 | hCoV-19/South Korea/KCDC2209/2020\|EPI_ISL_510548\|2 | .................... | .................... | .................... | .................... | .................... | .................... |
| F | 1959 | hCoV-19/South Korea/KCDC2211/2020\|EPI_ISL_510549\|2 | .................... | .................... | .................... | .................... | .................... | .................... |
| F | 1960 | hCoV-19/South Korea/KCDC2212/2020\|EPI_ISL_510550\|2 | .................... | .................... | .................... | .................... | .................... | .................... |
| F | 1961 | hCoV-19/South Korea/KCDC2213/2020\|EPI_ISL_510551\|2 | .................... | .................... | .................... | .................... | .................... | .................... |
| F | 1962 | hCoV-19/South Korea/KCDC2214/2020\|EPI_ISL_510552\|2 | .................... | .................... | .................... | .................... | .................... | .................... |
| F | 1963 | hCoV-19/South Korea/KCDC2215/2020\|EPI_ISL_510553\|2 | .................... | .................... | .................... | .................... | .................... | .................... |
| F | 1964 | hCoV-19/South Korea/KCDC2216/2020\|EPI_ISL_510554\|2 | .................... | .................... | .................... | .................... | .................... | .................... |
| F | 1965 | hCoV-19/South Korea/KCDC2217/2020\|EPI_ISL_510555\|2 | .................... | .................... | .................... | .................... | .................... | .................... |
| F | 1966 | hCoV-19/South Korea/KCDC2218/2020\|EPI_ISL_510556\|2 | .................... | .................... | .................... | .................... | .................... | .................... |
| F | 1967 | hCoV-19/South Korea/KCDC2219/2020\|EPI_ISL_510557\|2 | .................... | .................... | .................... | .................... | .................... | .................... |
| F | 1968 | hCoV-19/South Korea/KCDC2220/2020\|EPI_ISL_510558\|2 | .................... | .................... | .................... | .................... | .................... | .................... |
| F | 1969 | hCoV-19/South Korea/KCDC2221/2020\|EPI_ISL_510559\|2 | .................... | .................... | .................... | .................... | .................... | .................... |
| F | 1970 | hCoV-19/South Korea/KCDC2222/2020\|EPI_ISL_510560\|2 | .................... | .................... | .................... | .................... | .................... | .................... |
| F | 1971 | hCoV-19/South Korea/KCDC2223/2020\|EPI_ISL_510561\|2 | .................... | .................... | .................... | .................... | .................... | .................... |
| F | 1972 | hCoV-19/South Korea/KCDC2224/2020\|EPI_ISL_510562\|2 | .................... | .................... | .................... | .................... | .................... | .................... |
| F | 1973 | hCoV-19/South Korea/KCDC2225/2020\|EPI_ISL_510563\|2 | .................... | .................... | .................... | .................... | .................... | .................... |
| F | 1974 | hCoV-19/South Korea/KCDC2226/2020\|EPI_ISL_510564\|2 | .................... | .................... | .................... | .................... | .................... | .................... |
| F | 1975 | hCoV-19/South Korea/KCDC2227/2020\|EPI_ISL_510565\|2 | .................... | .................... | .................... | .................... | .................... | .................... |
| F | 1976 | hCoV-19/South Korea/KCDC2228/2020\|EPI_ISL_510566\|2 | .................... | .................... | .................... | .................... | .................... | .................... |
| F | 1977 | hCoV-19/South Korea/KCDC2232/2020\|EPI_ISL_510570\|2 | .................... | .................... | .................... | .................... | .................... | .................... |
| F | 1978 | hCoV-19/South Korea/KCDC2233/2020\|EPI_ISL_510571\|2 | .................... | .................... | .................... | .................... | .................... | .................... |
| F | 1979 | hCoV-19/South Korea/KCDC2234/2020\|EPI_ISL_510572\|2 | .................... | .................... | .................... | .................... | .................... | .................... |
| F | 1980 | hCoV-19/South Korea/KCDC2235/2020\|EPI_ISL_510573\|2 | .................... | .................... | .................... | .................... | .................... | .................... |
| F | 1981 | hCoV-19/South Korea/KCDC2239/2020\|EPI_ISL_510577\|2 | .................... | .................... | .................... | .................... | .................... | .................... |
| F | 1982 | hCoV-19/USA/WA-UW-10038/2020\|EPI_ISL_511854\|2020-0 | .................... | .................... | .................... | .................... | .................... | .................... |
| F | 1983 | hCoV-19/USA/WA-UW-10101/2020\|EPI_ISL_511855\|2020-0 | .................... | .................... | .................... | .................... | .................... | .................... |
| F | 1984 | hCoV-19/USA/WA-UW-10105/2020\|EPI_ISL_511856\|2020-0 | .................... | .................... | .................... | .................... | .................... | .................... |
| F | 1985 | hCoV-19/USA/WA-UW-10126/2020\|EPI_ISL_511860\|2020-0 | .................... | .................... | .................... | .................... | .................... | .................... |
| F | 1986 | hCoV-19/USA/WA-UW-10133/2020\|EPI_ISL_511861\|2020-0 | .................... | .................... | .................... | .................... | .................... | .................... |
| F | 1987 | hCoV-19/USA/CA-IGI-0066/2020\|EPI_ISL_511862\|2020-0 | .................... | .................... | .................... | .................... | .................... | .................... |
| F | 1988 | hCoV-19/USA/MD-HP00549/2020\|EPI_ISL_511869\|2020-04 | .................... | .................... | .................... | .................... | .................... | .................... |
| F | 1989 | hCoV-19/USA/MD-HP00567/2020\|EPI_ISL_511871\|2020-04 | .................... | .................... | .................... | .................... | .................... | .................... |
| F | 1990 | hCoV-19/USA/MD-HP01656/2020\|EPI_ISL_511874\|2020-04 | .................... | .................... | .................... | .................... | .................... | .................... |
| F | 1991 | hCoV-19/USA/MD-HP01661/2020\|EPI_ISL_511875\|2020-04 | .................... | .................... | .................... | .................... | .................... | .................... |
| F | 1992 | hCoV-19/Indonesia/JB-TFRIC19-L4173/2020\|EPI_ISL_51 | .................... | .................... | .................... | .................... | .................... | .................... |
| F | 1993 | hCoV-19/Vietnam/VNHN_0207/2020\|EPI_ISL_511891\|2020 | .................... | .................... | .................... | .................... | .................... | .................... |
| F | 1994 | hCoV-19/Vietnam/VNHN_0554/2020\|EPI_ISL_511892\|2020 | .................... | .................... | .................... | .................... | .................... | .................... |
| F | 1995 | hCoV-19/Vietnam/VNHN_0847/2020\|EPI_ISL_511893\|2020 | .................... | .................... | .................... | .................... | .................... | .................... |
| F | 1996 | hCoV-19/Vietnam/VNHN_0897/2020\|EPI_ISL_511894\|2020 | .................... | .................... | .................... | .................... | .................... | .................... |
| F | 1997 | hCoV-19/Vietnam/VNHN_0899/2020\|EPI_ISL_511895\|2020 | .................... | .................... | .................... | .................... | .................... | .................... |
| F | 1998 | hCoV-19/Vietnam/VNHN_1166/2020\|EPI_ISL_511896\|2020 | .................... | .................... | .................... | .................... | .................... | .................... |
| F | 1999 | hCoV-19/Vietnam/VNHN_1863/2020\|EPI_ISL_511897\|2020 | .................... | .................... | .................... | .................... | .................... | .................... |
| F | 2000 | hCoV-19/USA/NV-NSPHL-A0133/2020\|EPI_ISL_515401\|202 | .................... | .................... | .................... | .................... | .................... | .................... |
| F | 2001 | hCoV-19/USA/NV-NSPHL-A0136/2020\|EPI_ISL_515403\|202 | .................... | .................... | .................... | .................... | .................... | .................... |
| F | 2002 | hCoV-19/USA/NV-NSPHL-A0137/2020\|EPI_ISL_515404\|202 | .................... | .................... | .................... | .................... | .................... | .................... |
| F | 2003 | hCoV-19/USA/NV-NSPHL-A0138/2020\|EPI_ISL_515405\|202 | .................... | .................... | .................... | .................... | .................... | .................... |
| F | 2004 | hCoV-19/USA/NV-NSPHL-A0139/2020\|EPI_ISL_515406\|202 | .................... | .................... | .................... | .................... | .................... | .................... |
| F | 2005 | hCoV-19/USA/NV-NSPHL-A0141/2020\|EPI_ISL_515407\|202 | .................... | .................... | .................... | .................... | .................... | .................... |
| F | 2006 | hCoV-19/USA/NV-NSPHL-A0144/2020\|EPI_ISL_515409\|202 | .................... | .................... | .................... | .................... | .................... | .................... |
| F | 2007 | hCoV-19/USA/NV-NSPHL-A0145/2020\|EPI_ISL_515410\|202 | .................... | .................... | .................... | .................... | .................... | .................... |
| F | 2008 | hCoV-19/USA/NV-NSPHL-A0146/2020\|EPI_ISL_515411\|202 | .................... | .................... | .................... | .................... | .................... | .................... |
| F | 2009 | hCoV-19/USA/NV-NSPHL-A0147/2020\|EPI_ISL_515412\|202 | .................... | .A.................. | .................... | .................... | .................... | .................... |
| F | 2010 | hCoV-19/USA/NV-NSPHL-A0148/2020\|EPI_ISL_515413\|202 | .................... | .................... | .................... | .................... | .................... | .................... |
| F | 2011 | hCoV-19/USA/NV-NSPHL-A0149/2020\|EPI_ISL_515414\|202 | .................... | .................... | .................... | .................... | .................... | .................... |
| F | 2012 | hCoV-19/USA/NV-NSPHL-A0151/2020\|EPI_ISL_515415\|202 | .................... | .................... | .................... | .................... | .................... | .................... |
| F | 2013 | hCoV-19/USA/NV-NSPHL-A0152/2020\|EPI_ISL_515416\|202 | .................... | .................... | .................... | .................... | .................... | .................... |
| F | 2014 | hCoV-19/USA/NV-NSPHL-A0153/2020\|EPI_ISL_515417\|202 | .................... | .................... | .................... | .................... | .................... | .................... |

| | | | | | | | |
|---|---|---|---|---|---|---|---|
| F | 2015 hCoV-19/USA/VA-DCLS-0841/2020\|EPI_ISL_516708\|2020- | ···················· | ···················· | ···················· | ···················· | ···················· | ···················· |
| F | 2016 hCoV-19/USA/VA-DCLS-0843/2020\|EPI_ISL_516709\|2020- | ···················· | ···················· | ···················· | ···················· | ···················· | ···················· |
| F | 2017 hCoV-19/USA/VA-DCLS-0853/2020\|EPI_ISL_516716\|2020- | ···················· | ···················· | ···················· | ···················· | ···················· | ···················· |
| F | 2018 hCoV-19/USA/VA-DCLS-0854/2020\|EPI_ISL_516717\|2020- | ···················· | ···················· | ···················· | ···················· | ···················· | ···················· |
| F | 2019 hCoV-19/USA/VA-DCLS-0856/2020\|EPI_ISL_516718\|2020- | ···················· | ···A················ | ···················· | ···················· | ···················· | ···················· |
| F | 2020 hCoV-19/USA/VA-DCLS-0857/2020\|EPI_ISL_516719\|2020- | ···················· | ···················· | ···················· | ···················· | ···················· | ···················· |
| F | 2021 hCoV-19/USA/VA-DCLS-0858/2020\|EPI_ISL_516720\|2020- | ···················· | ···················· | ···················· | ···················· | ···················· | ···················· |
| F | 2022 hCoV-19/Peru/LIM-INS-053/2020\|EPI_ISL_516721\|2020- | ···················· | ···················· | ···················· | ···················· | ···················· | ···················· |
| F | 2023 hCoV-19/Peru/LIM-INS-054/2020\|EPI_ISL_516722\|2020- | ···················· | ···················· | ···················· | ···················· | ···················· | ···················· |
| F | 2024 hCoV-19/USA/CA-CZB-3648/2020\|EPI_ISL_516723\|2020-0 | ···················· | ·A·················· | ···················· | ···················· | ···················· | ···················· |
| F | 2025 hCoV-19/USA/CA-CZB-3649/2020\|EPI_ISL_516724\|2020-0 | ···················· | ·A·················· | ···················· | ···················· | ···················· | ···················· |
| F | 2026 hCoV-19/USA/CA-CZB-3650/2020\|EPI_ISL_516725\|2020-0 | ···················· | ·A·················· | ···················· | ···················· | ···················· | ···················· |
| F | 2027 hCoV-19/USA/CA-CZB-3651/2020\|EPI_ISL_516726\|2020-0 | ···················· | ·A·················· | ···················· | ···················· | ···················· | ···················· |
| F | 2028 hCoV-19/USA/CA-CZB-3652/2020\|EPI_ISL_516727\|2020-0 | ···················· | ···················· | ···················· | ···················· | ···················· | ···················· |
| F | 2029 hCoV-19/USA/CA-CZB-3653/2020\|EPI_ISL_516728\|2020-0 | ···················· | ···················· | ···················· | ···················· | ···················· | ···················· |
| F | 2030 hCoV-19/USA/CA-CZB-3654/2020\|EPI_ISL_516729\|2020-0 | ···················· | ···················· | ···················· | ···················· | ···················· | ···················· |
| F | 2031 hCoV-19/USA/CA-CZB-3655/2020\|EPI_ISL_516730\|2020-0 | ···················· | ·A·················· | ···················· | ···················· | ···················· | ···················· |
| F | 2032 hCoV-19/USA/CA-CZB-3656/2020\|EPI_ISL_516731\|2020-0 | ···················· | ·A·················· | ···················· | ···················· | ···················· | ···················· |
| F | 2033 hCoV-19/USA/CA-CZB-3657/2020\|EPI_ISL_516732\|2020-0 | ···················· | ·A·················· | ···················· | ···················· | ···················· | ···················· |
| F | 2034 hCoV-19/USA/CA-CZB-3658/2020\|EPI_ISL_516733\|2020-0 | ···················· | ·A·················· | ···················· | ···················· | ···················· | ···················· |
| F | 2035 hCoV-19/USA/CA-CZB-3659/2020\|EPI_ISL_516734\|2020-0 | ···················· | ·A·················· | ···················· | ···················· | ···················· | ···················· |
| F | 2036 hCoV-19/USA/CA-CZB-3660/2020\|EPI_ISL_516735\|2020-0 | ···················· | ·A·················· | ···················· | ···················· | ···················· | ···················· |
| F | 2037 hCoV-19/USA/CA-CZB-3661/2020\|EPI_ISL_516736\|2020-0 | ···················· | ·A·················· | ···················· | ···················· | ···················· | ···················· |
| F | 2038 hCoV-19/USA/CA-CZB-3663/2020\|EPI_ISL_516738\|2020-0 | ···················· | ·A·················· | ···················· | ···················· | ···················· | ···················· |
| F | 2039 hCoV-19/USA/CA-CZB-3664/2020\|EPI_ISL_516739\|2020-0 | ···················· | ·A·················· | ···················· | ···················· | ···················· | ···················· |
| F | 2040 hCoV-19/USA/CA-CZB-3665/2020\|EPI_ISL_516740\|2020-0 | ···················· | ·A·················· | ···················· | ···················· | ···················· | ···················· |
| F | 2041 hCoV-19/USA/CA-CZB-3666/2020\|EPI_ISL_516741\|2020-0 | ···················· | ·A·················· | ···················· | ···················· | ···················· | ···················· |
| F | 2042 hCoV-19/USA/CA-CZB-3667/2020\|EPI_ISL_516742\|2020-0 | ···················· | ·A·················· | ···················· | ···················· | ···················· | ···················· |
| F | 2043 hCoV-19/USA/CA-CZB-3668/2020\|EPI_ISL_516743\|2020-0 | ···················· | ·A·················· | ···················· | ···················· | ···················· | ···················· |
| F | 2044 hCoV-19/USA/CA-CZB-3669/2020\|EPI_ISL_516744\|2020-0 | ···················· | ·A·················· | ···················· | ···················· | ···················· | ···················· |
| F | 2045 hCoV-19/USA/CA-CZB-3670/2020\|EPI_ISL_516745\|2020-0 | ···················· | ·A·················· | ···················· | ···················· | ···················· | ···················· |
| F | 2046 hCoV-19/USA/CA-CZB-3671/2020\|EPI_ISL_516746\|2020-0 | ···················· | ·A·················· | ···················· | ···················· | ···················· | ···················· |
| F | 2047 hCoV-19/USA/CA-CZB-3672/2020\|EPI_ISL_516747\|2020-0 | ···················· | ·A·················· | ···················· | ···················· | ···················· | ···················· |
| F | 2048 hCoV-19/USA/SC-20-0546-T00/2020\|EPI_ISL_516748\|202 | ···················· | ···················· | ···················· | ···················· | ···················· | ···················· |
| F | 2050 hCoV-19/England/LIVE-A58E7/2020\|EPI_ISL_499645\|202 | ···················· | ···················· | ···················· | ···················· | ···················· | ···················· |
| F | 2051 hCoV-19/England/LIVE-A58F6/2020\|EPI_ISL_499646\|202 | ···················· | ···················· | ···················· | ···················· | ···················· | ···················· |
| F | 2053 hCoV-19/England/LIVE-A59A8/2020\|EPI_ISL_499651\|202 | ···················· | ···················· | ···················· | ···················· | ···················· | ···················· |
| F | 2054 hCoV-19/England/LIVE-A5A5A/2020\|EPI_ISL_499653\|202 | ···················· | ···················· | ···················· | ···················· | ···················· | ···················· |
| F | 2055 hCoV-19/England/LIVE-A5A78/2020\|EPI_ISL_499654\|202 | ···················· | ···················· | ···················· | ···················· | ···················· | ···················· |
| F | 2056 hCoV-19/India/KA-IB12/2020\|EPI_ISL_508290\|2020-06- | ···················· | ···················· | ···················· | ···················· | ···················· | ···················· |
| F | 2057 hCoV-19/India/KA-IB13/2020\|EPI_ISL_508291\|2020-06- | ···················· | ···················· | ···················· | ···················· | ···················· | ···················· |
| F | 2058 hCoV-19/India/KA-IB14/2020\|EPI_ISL_508292\|2020-06- | ···················· | ···················· | ··T················· | ···················· | ···················· | ···················· |
| F | 2059 hCoV-19/India/KA-IB15/2020\|EPI_ISL_508293\|2020-06- | ···················· | ···················· | ···················· | ···················· | ···················· | ···················· |
| F | 2060 hCoV-19/India/KA-IB16/2020\|EPI_ISL_508294\|2020-06- | ···················· | ···················· | ···················· | ···················· | ···················· | ···················· |
| F | 2061 hCoV-19/India/KA-IB17/2020\|EPI_ISL_508295\|2020-06- | ···················· | ···················· | ···················· | ···················· | ···················· | ···················· |
| F | 2062 hCoV-19/India/KA-IB18/2020\|EPI_ISL_508296\|2020-06- | ···················· | ···················· | ···················· | ···················· | ···················· | ···················· |
| F | 2063 hCoV-19/India/KA-IB19/2020\|EPI_ISL_508297\|2020-06- | ···················· | ···················· | ···················· | ···················· | ···················· | ···················· |
| F | 2064 hCoV-19/India/KA-IB2/2020\|EPI_ISL_508298\|2020-06-2 | ···················· | ···················· | ···················· | ···················· | ···················· | ···················· |
| F | 2065 hCoV-19/India/KA-IB21/2020\|EPI_ISL_508299\|2020-06- | ···················· | ···················· | ···················· | ···················· | ···················· | ···················· |
| F | 2066 hCoV-19/India/KA-IB26/2020\|EPI_ISL_508304\|2020-06- | ···················· | ···················· | ···················· | ···················· | ···················· | ···················· |
| F | 2067 hCoV-19/India/KA-IB27/2020\|EPI_ISL_508305\|2020-05- | ···················· | ···················· | ···················· | ···················· | ···················· | ···················· |
| F | 2068 hCoV-19/India/KA-IB29/2020\|EPI_ISL_508307\|2020-06- | ···················· | ···················· | ···················· | ···················· | ···················· | ···················· |
| F | 2069 hCoV-19/India/KA-IB31/2020\|EPI_ISL_508310\|2020-06- | ···················· | ···················· | ···················· | ···················· | ···················· | ···················· |
| F | 2070 hCoV-19/India/KA-IB32/2020\|EPI_ISL_508311\|2020-06- | ···················· | ···················· | ···················· | ···················· | ···················· | ···················· |
| F | 2071 hCoV-19/India/KA-IB33/2020\|EPI_ISL_508312\|2020-06- | ···················· | ···················· | ···················· | ···················· | ···················· | ···················· |
| F | 2072 hCoV-19/India/KA-IB34/2020\|EPI_ISL_508313\|2020-06- | ···················· | ···················· | ···················· | ···················· | ···················· | ···················· |
| F | 2073 hCoV-19/India/KA-IB36/2020\|EPI_ISL_508315\|2020-06- | ···················· | ···················· | ···················· | ···················· | ···················· | ···················· |
| F | 2074 hCoV-19/India/KA-IB38/2020\|EPI_ISL_508316\|2020-06- | ···················· | ···················· | ···················· | ···················· | ···················· | ···················· |
| F | 2075 hCoV-19/India/KA-IB39/2020\|EPI_ISL_508317\|2020-06- | ···················· | ···················· | ···················· | ···················· | ···················· | ···················· |
| F | 2076 hCoV-19/India/KA-IB4/2020\|EPI_ISL_508318\|2020-06-1 | ···················· | ···················· | ···················· | ···················· | ···················· | ···················· |
| F | 2077 hCoV-19/India/KA-IB40/2020\|EPI_ISL_508319\|2020-06- | ···················· | ···················· | ···················· | ···················· | ···················· | ···················· |
| F | 2078 hCoV-19/India/KA-IB41/2020\|EPI_ISL_508320\|2020-06- | ···················· | ···················· | ···················· | ···················· | ···················· | ···················· |
| F | 2079 hCoV-19/India/KA-IB42/2020\|EPI_ISL_508321\|2020-06- | ···················· | ···················· | ···················· | ···················· | ···················· | ···················· |
| F | 2080 hCoV-19/India/KA-IB43/2020\|EPI_ISL_508322\|2020-06- | ···················· | ···················· | ···················· | ···················· | ···················· | ···················· |
| F | 2081 hCoV-19/India/KA-IB44/2020\|EPI_ISL_508323\|2020-06- | ···················· | ···················· | ···················· | ···················· | ···················· | ···················· |
| F | 2082 hCoV-19/India/KA-IB46/2020\|EPI_ISL_508324\|2020-05- | ···················· | ·A·················· | ···················· | ···················· | ···················· | ··A················· |
| F | 2083 hCoV-19/India/KA-IB47/2020\|EPI_ISL_508325\|2020-06- | ···················· | ···················· | ···················· | ···················· | ···················· | ···················· |
| F | 2084 hCoV-19/India/KA-IB48/2020\|EPI_ISL_508326\|2020-06- | ···················· | ···················· | ···················· | ···················· | ···················· | ···················· |
| F | 2085 hCoV-19/India/KA-IB5/2020\|EPI_ISL_508327\|2020-06-0 | ···················· | ···················· | ···················· | ···················· | ···················· | ···················· |
| F | 2086 hCoV-19/India/KA-IB50/2020\|EPI_ISL_508328\|2020-05- | ···················· | ···················· | ···················· | ···················· | ···················· | ···················· |
| F | 2087 hCoV-19/India/KA-IB51/2020\|EPI_ISL_508329\|2020-06- | ···················· | ···················· | ···················· | ···················· | ···················· | ···················· |
| F | 2088 hCoV-19/Spain/AN-IBV-001981/2020\|EPI_ISL_509620\|20 | ···················· | ···················· | ···················· | ···················· | ···················· | ···················· |
| F | 2089 hCoV-19/Spain/AN-IBV-001984/2020\|EPI_ISL_509621\|20 | ···················· | ···················· | ···················· | ···················· | ···················· | ···················· |
| F | 2090 hCoV-19/Spain/AN-IBV-001985/2020\|EPI_ISL_509622\|20 | ···················· | ···················· | ···················· | ···················· | ···················· | ···················· |
| F | 2091 hCoV-19/Spain/AN-IBV-001987/2020\|EPI_ISL_509623\|20 | ···················· | ···················· | ···················· | ···················· | ··NN················ | ···················· |
| F | 2092 hCoV-19/Spain/AN-IBV-001989/2020\|EPI_ISL_509624\|20 | ···················· | ···················· | ···················· | ···················· | ···················· | ···················· |
| F | 2093 hCoV-19/Spain/AN-IBV-001995/2020\|EPI_ISL_509625\|20 | ···················· | ···················· | ···················· | ···················· | ···················· | ···················· |
| F | 2094 hCoV-19/Spain/AN-IBV-001996/2020\|EPI_ISL_509626\|20 | ···················· | ···················· | ···················· | ···················· | ···················· | ···················· |
| F | 2095 hCoV-19/Spain/AN-IBV-002016/2020\|EPI_ISL_509627\|20 | ···················· | ···················· | ···················· | ···················· | ···················· | ···················· |
| F | 2096 hCoV-19/Spain/AN-IBV-002017/2020\|EPI_ISL_509628\|20 | ···················· | ···················· | ···················· | ···················· | ···················· | ···················· |

| | | | | | | | | |
|---|---|---|---|---|---|---|---|---|
| F | 2097 | hCoV-19/Spain/AN-IBV-002019/2020\|EPI_ISL_509629\|20 | ···················· | ···················· | ···················· | ···················· | ···················· | ···················· |
| F | 2098 | hCoV-19/Spain/PV-IBV-005143/2020\|EPI_ISL_509635\|20 | ···················· | ···················· | ···················· | ···················· | ···················· | ···················· |
| F | 2099 | hCoV-19/Spain/PV-IBV-005145/2020\|EPI_ISL_509636\|20 | ···················· | ···················· | ···················· | ···················· | ···················· | ···················· |
| F | 2100 | hCoV-19/Spain/PV-IBV-005147/2020\|EPI_ISL_509637\|20 | ···················· | ···················· | ···················· | ···················· | ···················· | ···················· |
| F | 2101 | hCoV-19/Spain/PV-IBV-005148/2020\|EPI_ISL_509638\|20 | ···················· | ···················· | ···················· | ···················· | ···················· | ···················· |
| F | 2102 | hCoV-19/Spain/PV-IBV-005150/2020\|EPI_ISL_509640\|20 | ···················· | ···················· | ···················· | ···················· | ···················· | ···················· |
| F | 2103 | hCoV-19/Spain/PV-IBV-005153/2020\|EPI_ISL_509641\|20 | ···················· | ···················· | ···················· | ···················· | ···················· | ···················· |
| F | 2104 | hCoV-19/Spain/PV-IBV-005154/2020\|EPI_ISL_509642\|20 | ···················· | ···················· | ···················· | ···················· | ···················· | ···················· |
| F | 2106 | hCoV-19/Spain/PV-IBV-005157/2020\|EPI_ISL_509644\|20 | ···················· | ···················· | ···················· | ···················· | ···················· | ···················· |
| F | 2107 | hCoV-19/Spain/PV-IBV-005158/2020\|EPI_ISL_509645\|20 | ···················· | ···················· | ···················· | ···················· | ···················· | ···················· |
| F | 2108 | hCoV-19/Spain/PV-IBV-005159/2020\|EPI_ISL_509646\|20 | ···················· | ···················· | ···················· | ···················· | ···················· | ···················· |
| F | 2109 | hCoV-19/Spain/PV-IBV-005161/2020\|EPI_ISL_509647\|20 | ···················· | ···················· | ···················· | ···················· | ···················· | ···················· |
| F | 2110 | hCoV-19/Spain/PV-IBV-005163/2020\|EPI_ISL_509648\|20 | ···················· | ···················· | ···················· | ···················· | ···················· | ···················· |
| F | 2111 | hCoV-19/Spain/PV-IBV-005168/2020\|EPI_ISL_509649\|20 | ···················· | ···················· | ···················· | ···················· | ···················· | ···················· |
| F | 2112 | hCoV-19/Spain/PV-IBV-005169/2020\|EPI_ISL_509650\|20 | ···················· | ···················· | ···················· | ···················· | ···················· | ···················· |
| F | 2113 | hCoV-19/Spain/PV-IBV-005172/2020\|EPI_ISL_509651\|20 | ···················· | ···················· | ···················· | ···················· | ···················· | ···················· |
| F | 2114 | hCoV-19/Spain/PV-IBV-005173/2020\|EPI_ISL_509652\|20 | ···················· | ···················· | ···················· | ···················· | ···················· | ···················· |
| F | 2115 | hCoV-19/Spain/PV-IBV-005174/2020\|EPI_ISL_509653\|20 | ···················· | ···················· | ···················· | ···················· | ···················· | ···················· |
| F | 2116 | hCoV-19/Spain/PV-IBV-005175/2020\|EPI_ISL_509654\|20 | ···················· | ···················· | ···················· | ···················· | ···················· | ···················· |
| F | 2117 | hCoV-19/South Korea/KCDC2266/2020\|EPI_ISL_510604\|2 | ···················· | ···················· | ···················· | ···················· | ···················· | ···················· |
| F | 2118 | hCoV-19/South Korea/KCDC2267/2020\|EPI_ISL_510605\|2 | ···················· | ···················· | ···················· | ···················· | ···················· | ···················· |
| F | 2119 | hCoV-19/South Korea/KCDC2268/2020\|EPI_ISL_510606\|2 | ···················· | ···················· | ···················· | ···················· | ···················· | ···················· |
| F | 2120 | hCoV-19/South Korea/KCDC2269/2020\|EPI_ISL_510607\|2 | ···················· | ···················· | ···················· | ···················· | ···················· | ···················· |
| F | 2121 | hCoV-19/South Korea/KCDC2274/2020\|EPI_ISL_510611\|2 | ···················· | ···················· | ···················· | ···················· | ···················· | ···················· |
| F | 2122 | hCoV-19/South Korea/KCDC2275/2020\|EPI_ISL_510612\|2 | ···················· | ···················· | ···················· | ···················· | ···················· | ···················· |
| F | 2123 | hCoV-19/South Korea/KCDC2276/2020\|EPI_ISL_510613\|2 | ···················· | ···················· | ···················· | ···················· | ···················· | ···················· |
| F | 2124 | hCoV-19/South Korea/KCDC2277/2020\|EPI_ISL_510614\|2 | ···················· | ···················· | ···················· | ···················· | ···················· | ···················· |
| F | 2125 | hCoV-19/South Korea/KCDC2278/2020\|EPI_ISL_510615\|2 | ···················· | ···················· | ···················· | ···················· | ···················· | ···················· |
| F | 2126 | hCoV-19/South Korea/KCDC2279/2020\|EPI_ISL_510616\|2 | ···················· | ···················· | ···················· | ···················· | ···················· | ···················· |
| F | 2127 | hCoV-19/South Korea/KCDC2280/2020\|EPI_ISL_510617\|2 | ···················· | ···················· | ···················· | ···················· | ···················· | ···················· |
| F | 2128 | hCoV-19/South Korea/KCDC2281/2020\|EPI_ISL_510618\|2 | ···················· | ···················· | ···················· | ···················· | ···················· | ···················· |
| F | 2129 | hCoV-19/South Korea/KCDC2282/2020\|EPI_ISL_510619\|2 | ···················· | ···················· | ···················· | ···················· | ···················· | ···················· |
| F | 2130 | hCoV-19/South Korea/KCDC2283/2020\|EPI_ISL_510620\|2 | ···················· | ···················· | ···················· | ···················· | ···················· | ···················· |
| F | 2131 | hCoV-19/South Korea/KCDC2284/2020\|EPI_ISL_510621\|2 | ···················· | ···················· | ···················· | ···················· | ···················· | ···················· |
| F | 2132 | hCoV-19/South Korea/KCDC2285/2020\|EPI_ISL_510622\|2 | ···················· | ···················· | ···················· | ···················· | ···················· | ···················· |
| F | 2133 | hCoV-19/South Korea/KCDC2286/2020\|EPI_ISL_510623\|2 | ···················· | ···················· | ···················· | ···················· | ···················· | ···················· |
| F | 2134 | hCoV-19/South Korea/KCDC2287/2020\|EPI_ISL_510624\|2 | ···················· | ···················· | ···················· | ···················· | ···················· | ···················· |
| F | 2135 | hCoV-19/South Korea/KCDC2288/2020\|EPI_ISL_510625\|2 | ···················· | ···················· | ···················· | ···················· | ···················· | ···················· |
| F | 2136 | hCoV-19/South Korea/KCDC2289/2020\|EPI_ISL_510626\|2 | ···················· | ···················· | ···················· | ···················· | ···················· | ···················· |
| F | 2137 | hCoV-19/South Korea/KCDC2291/2020\|EPI_ISL_510627\|2 | ···················· | ···················· | ···················· | ···················· | ···················· | ···················· |
| F | 2138 | hCoV-19/South Korea/KCDC2292/2020\|EPI_ISL_510628\|2 | ···················· | ···················· | ···················· | ···················· | ···················· | ···················· |
| F | 2139 | hCoV-19/South Korea/KCDC2293/2020\|EPI_ISL_510629\|2 | ···················· | ···················· | ···················· | ···················· | ···················· | ···················· |
| F | 2140 | hCoV-19/South Korea/KCDC2294/2020\|EPI_ISL_510630\|2 | ···················· | ···················· | ···················· | ···················· | ···················· | ···················· |
| F | 2141 | hCoV-19/South Korea/KCDC2295/2020\|EPI_ISL_510631\|2 | ···················· | ···················· | ···················· | ···················· | ···················· | ···················· |
| F | 2142 | hCoV-19/South Korea/KCDC2296/2020\|EPI_ISL_510632\|2 | ···················· | ···················· | ···················· | ···················· | ···················· | ···················· |
| F | 2143 | hCoV-19/South Korea/KCDC2297/2020\|EPI_ISL_510633\|2 | ···················· | ···················· | ···················· | ···················· | ···················· | ···················· |
| F | 2144 | hCoV-19/South Korea/KCDC2298/2020\|EPI_ISL_510634\|2 | ···················· | ···················· | ···················· | ···················· | ···················· | ···················· |
| F | 2145 | hCoV-19/South Korea/KCDC2299/2020\|EPI_ISL_510635\|2 | ···················· | ···················· | ···················· | ···················· | ···················· | ···················· |
| F | 2146 | hCoV-19/South Korea/KCDC2300/2020\|EPI_ISL_510636\|2 | ···················· | ···················· | ···················· | ···················· | ···················· | ···················· |
| F | 2147 | hCoV-19/South Korea/KCDC2301/2020\|EPI_ISL_510637\|2 | ···················· | ···················· | ···················· | ···················· | ···················· | ···················· |
| F | 2148 | hCoV-19/South Korea/KCDC2302/2020\|EPI_ISL_510638\|2 | ···················· | ···················· | ···················· | ···················· | ···················· | ···················· |
| F | 2149 | hCoV-19/South Korea/KCDC2303/2020\|EPI_ISL_510639\|2 | ···················· | ···················· | ···················· | ···················· | ···················· | ···················· |
| F | 2150 | hCoV-19/South Korea/KCDC2304/2020\|EPI_ISL_510640\|2 | ···················· | ···················· | ···················· | ···················· | ···················· | ···················· |
| F | 2151 | hCoV-19/South Korea/KCDC2305/2020\|EPI_ISL_510641\|2 | ···················· | ···················· | ···················· | ···················· | ···················· | ···················· |
| F | 2152 | hCoV-19/South Korea/KCDC2306/2020\|EPI_ISL_510642\|2 | ···················· | ···················· | ···················· | ···················· | ···················· | ···················· |
| F | 2153 | hCoV-19/South Korea/KCDC2307/2020\|EPI_ISL_510643\|2 | ···················· | ···················· | ···················· | ···················· | ···················· | ···················· |
| F | 2154 | hCoV-19/India/MH-GA90/2020\|EPI_ISL_511935\|2020-06- | ···················· | ···················· | ···················· | ···················· | ···················· | ···················· |
| F | 2155 | hCoV-19/India/HR-TF19/2020\|EPI_ISL_511943\|2020-06- | ···················· | ·A·················· | ···················· | ···················· | ···················· | ··A················· |
| F | 2156 | hCoV-19/India/HR-TF21/2020\|EPI_ISL_511944\|2020-06- | ···················· | ·A·················· | ···················· | ···················· | ···················· | ··A················· |
| F | 2157 | hCoV-19/England/BRIS-13308B/2020\|EPI_ISL_511955\|20 | ···················· | ·············A······ | ···················· | ···················· | ···················· | ···················· |
| F | 2158 | hCoV-19/England/BRIS-132F16/2020\|EPI_ISL_511956\|20 | ···················· | ···················· | ···················· | ···················· | ···················· | ···················· |
| F | 2159 | hCoV-19/England/BRIS-13312E/2020\|EPI_ISL_511957\|20 | ···················· | ···················· | ···················· | ···················· | ···················· | ···················· |
| F | 2160 | hCoV-19/England/BRIS-132F8F/2020\|EPI_ISL_511958\|20 | ···················· | ···················· | ···················· | ···················· | ···················· | ···················· |
| F | 2161 | hCoV-19/England/BRIS-132DEF/2020\|EPI_ISL_511960\|20 | ···················· | ···················· | ···················· | ···················· | ···················· | ···················· |
| F | 2162 | hCoV-19/England/BRIS-132FAD/2020\|EPI_ISL_511964\|20 | ···················· | ···················· | ···················· | ···················· | ···················· | ···················· |
| F | 2163 | hCoV-19/England/BRIS-132DA3/2020\|EPI_ISL_511966\|20 | ···················· | ···················· | ···················· | ···················· | ·········T·········· | ···················· |
| F | 2164 | hCoV-19/England/BRIS-13267B/2020\|EPI_ISL_511967\|20 | ···················· | ···················· | ···················· | ···················· | ···················· | ···················· |
| F | 2165 | hCoV-19/England/BRIS-132D2B/2020\|EPI_ISL_511968\|20 | ···················· | ···················· | ···················· | ···················· | ···················· | ···················· |
| F | 2166 | hCoV-19/England/CAMB-1B385E/2020\|EPI_ISL_511970\|20 | ···················· | ···················· | ···················· | ···················· | ···················· | ···················· |
| F | 2167 | hCoV-19/England/BRIS-13314C/2020\|EPI_ISL_511971\|20 | ···················· | ···················· | ···················· | ···················· | ···················· | ···················· |
| F | 2168 | hCoV-19/England/BRIS-132D85/2020\|EPI_ISL_511972\|20 | ···················· | ···················· | ···················· | ···················· | ·········T·········· | ···················· |
| F | 2169 | hCoV-19/England/BRIS-133012/2020\|EPI_ISL_511973\|20 | ···················· | ·············A······ | ···················· | ···················· | ···················· | ···················· |
| F | 2170 | hCoV-19/England/BRIS-132CA6/2020\|EPI_ISL_511974\|20 | ···················· | ···················· | ···················· | ···················· | ···················· | ···················· |
| F | 2171 | hCoV-19/DRC/4279/2020\|EPI_ISL_514114\|2020-04-28 | ···················· | ···················· | ···················· | ···················· | ···················· | ···················· |
| F | 2172 | hCoV-19/DRC/4284/2020\|EPI_ISL_514115\|2020-04-28 | ···················· | ···················· | ···················· | ···················· | ···················· | ···················· |
| F | 2173 | hCoV-19/DRC/4690/2020\|EPI_ISL_514117\|2020-04-30 | ···················· | ···················· | ···················· | ···················· | ···················· | ···················· |
| F | 2174 | hCoV-19/DRC/4693/2020\|EPI_ISL_514118\|2020-04-30 | ···················· | ···················· | ···················· | ···················· | ···················· | ···················· |
| F | 2175 | hCoV-19/DRC/4785/2020\|EPI_ISL_514119\|2020-05-01 | ···················· | ···················· | ···················· | ···················· | ···················· | ···················· |
| F | 2176 | hCoV-19/DRC/7211/2020\|EPI_ISL_514122\|2020-05-13 | ···················· | ···················· | ···················· | ···················· | ···················· | ···················· |
| F | 2177 | hCoV-19/USA/UN-CDC-7650/2020\|EPI_ISL_514128\|2020-0 | ···················· | ···················· | ···················· | ···················· | ···················· | ···················· |

| | | | | | | | | |
|---|---|---|---|---|---|---|---|---|
| F | 2178 | hCoV-19/Bangladesh/BCSIR-NILMRC-356/2020\|EPI_ISL_5 | ···················· | ···················· | ···················· | ···················· | ···················· | ···················· |
| F | 2179 | hCoV-19/Bangladesh/BCSIR-NILMRC-358/2020\|EPI_ISL_5 | ···················· | ···················· | ···················· | ···················· | ···················· | ···················· |
| F | 2180 | hCoV-19/Brazil/RO-01/2020\|EPI_ISL_514131\|2020-05-1 | ···················· | ···················· | ···················· | ···················· | ···················· | ···················· |
| F | 2181 | hCoV-19/Brazil/RO-02/2020\|EPI_ISL_514132\|2020-05-1 | ···················· | ···················· | ···················· | ···················· | ···················· | ···················· |
| F | 2182 | hCoV-19/Brazil/RO-03/2020\|EPI_ISL_514133\|2020-05-1 | ···················· | ···················· | ···················· | ···················· | ···················· | ···················· |
| F | 2183 | hCoV-19/Brazil/RO-04/2020\|EPI_ISL_514134\|2020-05-1 | ···················· | ···················· | ···················· | ···················· | ···················· | ···················· |
| F | 2184 | hCoV-19/Brazil/RO-05/2020\|EPI_ISL_514135\|2020-05-0 | ···················· | ···················· | ···················· | ···················· | ···················· | ···················· |
| F | 2185 | hCoV-19/Brazil/RO-06/2020\|EPI_ISL_514136\|2020-05-0 | ···················· | ···················· | ···················· | ···················· | ···················· | ···················· |
| F | 2186 | hCoV-19/Brazil/RO-07/2020\|EPI_ISL_514137\|2020-05-0 | ···················· | ···················· | ···················· | ···················· | ···················· | ···················· |
| F | 2187 | hCoV-19/Brazil/RO-08/2020\|EPI_ISL_514138\|2020-06-0 | ···················· | ···················· | ···················· | ···················· | ···················· | ···················· |
| F | 2188 | hCoV-19/USA/FL-BPHL-0794/2020\|EPI_ISL_514140\|2020- | ···················· | ···················· | ···················· | ···················· | ···················· | ···················· |
| F | 2191 | hCoV-19/USA/FL-BPHL-0797/2020\|EPI_ISL_514143\|2020- | ···················· | ···················· | ···················· | ···················· | ···················· | ···················· |
| F | 2193 | hCoV-19/USA/FL-BPHL-0799/2020\|EPI_ISL_514145\|2020- | ···················· | ···················· | ···················· | ···················· | ···················· | ···················· |
| F | 2194 | hCoV-19/USA/FL-BPHL-0800/2020\|EPI_ISL_514146\|2020- | ···················· | ···················· | ···················· | ···················· | ···················· | ···················· |
| F | 2198 | hCoV-19/USA/FL-BPHL-0806/2020\|EPI_ISL_514152\|2020- | ···················· | ···················· | ···················· | ···················· | ···················· | ···················· |
| F | 2199 | hCoV-19/USA/FL-BPHL-0807/2020\|EPI_ISL_514153\|2020- | ···················· | ···················· | ···················· | ···················· | ···················· | ···················· |
| F | 2200 | hCoV-19/USA/FL-BPHL-0809/2020\|EPI_ISL_514155\|2020- | ···················· | ···················· | ···················· | ···················· | ···················· | ···················· |
| F | 2201 | hCoV-19/USA/FL-BPHL-0810/2020\|EPI_ISL_514156\|2020- | ···················· | ···················· | ···················· | ···················· | ···················· | ···················· |
| F | 2202 | hCoV-19/USA/FL-BPHL-0811/2020\|EPI_ISL_514157\|2020- | ···················· | ···················· | ···················· | ···················· | ···················· | ···················· |
| F | 2204 | hCoV-19/USA/FL-BPHL-0813/2020\|EPI_ISL_514159\|2020- | ···················· | ···················· | ···················· | ···················· | ···················· | ···················· |
| F | 2205 | hCoV-19/USA/FL-BPHL-0814/2020\|EPI_ISL_514160\|2020- | ···················· | ···················· | ···················· | ···················· | ···················· | ···················· |
| F | 2206 | hCoV-19/USA/FL-BPHL-0815/2020\|EPI_ISL_514161\|2020- | ···················· | ···················· | ···················· | ···················· | ···················· | ···················· |
| F | 2207 | hCoV-19/USA/FL-BPHL-0816/2020\|EPI_ISL_514162\|2020- | ···················· | ···················· | ···················· | ···················· | ···················· | ···················· |
| F | 2208 | hCoV-19/USA/FL-BPHL-0817/2020\|EPI_ISL_514163\|2020- | ···················· | ···················· | ···················· | ···················· | ···················· | ···················· |
| F | 2209 | hCoV-19/USA/FL-BPHL-0822/2020\|EPI_ISL_514168\|2020- | ···················· | ···················· | ···················· | ···················· | ···················· | ···················· |
| F | 2210 | hCoV-19/USA/FL-BPHL-0823/2020\|EPI_ISL_514169\|2020- | ···················· | ···················· | ···················· | ···················· | ···················· | ···················· |
| F | 2211 | hCoV-19/USA/NV-NSPHL-A0154/2020\|EPI_ISL_515418\|202 | ···················· | ···················· | ···················· | ···················· | ···················· | ···················· |
| F | 2212 | hCoV-19/USA/NV-NSPHL-A0155/2020\|EPI_ISL_515419\|202 | ···················· | ···················· | ···················· | ···················· | ···················· | ···················· |
| F | 2213 | hCoV-19/USA/NV-NSPHL-A0156/2020\|EPI_ISL_515420\|202 | ···················· | ···················· | ···················· | ···················· | ···················· | ···················· |
| F | 2214 | hCoV-19/USA/NV-NSPHL-A0157/2020\|EPI_ISL_515421\|202 | ···················· | ···················· | ···················· | ···················· | ···················· | ···················· |
| F | 2215 | hCoV-19/USA/NV-NSPHL-A0160/2020\|EPI_ISL_515422\|202 | ···················· | ···················· | ···················· | ···················· | ···················· | ···················· |
| F | 2216 | hCoV-19/USA/NV-NSPHL-A0161/2020\|EPI_ISL_515423\|202 | ···················· | ···················· | ···················· | ···················· | ···················· | ···················· |
| F | 2217 | hCoV-19/USA/NV-NSPHL-A0162/2020\|EPI_ISL_515424\|202 | ···················· | ···················· | ···················· | ···················· | ···················· | ···················· |
| F | 2218 | hCoV-19/USA/NV-NSPHL-A0163/2020\|EPI_ISL_515425\|202 | ···················· | ···················· | ···················· | ···················· | ···················· | ···················· |
| F | 2219 | hCoV-19/USA/NV-NSPHL-A0164/2020\|EPI_ISL_515426\|202 | ···················· | ···················· | ···················· | ···················· | ···················· | ···················· |
| F | 2220 | hCoV-19/USA/NV-NSPHL-A0165/2020\|EPI_ISL_515427\|202 | ···················· | ···················· | ···················· | ···················· | ···················· | ···················· |
| F | 2221 | hCoV-19/USA/NV-NSPHL-A0167/2020\|EPI_ISL_515428\|202 | ···················· | ···················· | ···················· | ···················· | ···················· | ···················· |
| F | 2222 | hCoV-19/USA/NV-NSPHL-A0171/2020\|EPI_ISL_515429\|202 | ···················· | ···················· | ···················· | ···················· | ···················· | ···················· |
| F | 2223 | hCoV-19/USA/NV-NSPHL-A0172/2020\|EPI_ISL_515430\|202 | ···················· | ···················· | ···················· | ···················· | ···················· | ···················· |
| F | 2224 | hCoV-19/USA/NV-NSPHL-A0173/2020\|EPI_ISL_515431\|202 | ···················· | ···················· | ···················· | ···················· | ···················· | ···················· |
| F | 2225 | hCoV-19/USA/NV-NSPHL-A0174/2020\|EPI_ISL_515432\|202 | ···················· | ···················· | ···················· | ···················· | ···················· | ···················· |
| F | 2226 | hCoV-19/USA/NV-NSPHL-A0175/2020\|EPI_ISL_515433\|202 | ···················· | ···················· | ···················· | ···················· | ···················· | ···················· |
| F | 2227 | hCoV-19/USA/NV-NSPHL-A0176/2020\|EPI_ISL_515434\|202 | ···················· | ···················· | ···················· | ···················· | ···················· | ···················· |
| F | 2228 | hCoV-19/USA/NV-NSPHL-A0178/2020\|EPI_ISL_515436\|202 | ···················· | ···················· | ···················· | ···················· | ···················· | ···················· |
| F | 2229 | hCoV-19/USA/NV-NSPHL-A0180/2020\|EPI_ISL_515437\|202 | ···················· | ···················· | ···················· | ···················· | ···················· | ···················· |
| F | 2230 | hCoV-19/USA/NV-NSPHL-A0181/2020\|EPI_ISL_515438\|202 | ···················· | ···················· | ···················· | ···················· | ···················· | ···················· |
| F | 2231 | hCoV-19/USA/NV-NSPHL-A0182/2020\|EPI_ISL_515439\|202 | ···················· | ···················· | ···················· | ···················· | ···················· | ···················· |
| F | 2232 | hCoV-19/USA/NV-NSPHL-A0183/2020\|EPI_ISL_515440\|202 | ···················· | ···················· | ···················· | ···················· | ···················· | ···················· |
| F | 2233 | hCoV-19/USA/NV-NSPHL-A0185/2020\|EPI_ISL_515441\|202 | ···················· | ···················· | ···················· | ···················· | ···················· | ···················· |
| F | 2234 | hCoV-19/USA/NV-NSPHL-A0186/2020\|EPI_ISL_515442\|202 | ···················· | ···················· | ···················· | ···················· | ···················· | ···················· |
| F | 2235 | hCoV-19/USA/NV-NSPHL-A0187/2020\|EPI_ISL_515443\|202 | ···················· | ···················· | ···················· | ···················· | ···················· | ···················· |
| F | 2236 | hCoV-19/USA/NV-NSPHL-A0188/2020\|EPI_ISL_515444\|202 | ···················· | ···················· | ···················· | ···················· | ···················· | ···················· |
| F | 2237 | hCoV-19/USA/NV-NSPHL-A0189/2020\|EPI_ISL_515445\|202 | ···················· | ···················· | ···················· | ···················· | ···················· | ···················· |
| F | 2238 | hCoV-19/USA/NV-NSPHL-A0190/2020\|EPI_ISL_515446\|202 | ···················· | ···················· | ···················· | ···················· | ···················· | ···················· |
| F | 2239 | hCoV-19/USA/NV-NSPHL-A0193/2020\|EPI_ISL_515447\|202 | ···················· | ···················· | ···················· | ···················· | ···················· | ···················· |
| F | 2240 | hCoV-19/USA/NV-NSPHL-A0194/2020\|EPI_ISL_515448\|202 | ···················· | ···················· | ···················· | ···················· | ···················· | ···················· |
| F | 2241 | hCoV-19/USA/NV-NSPHL-A0195/2020\|EPI_ISL_515449\|202 | ···················· | ···················· | ···················· | ···················· | ···················· | ···················· |
| F | 2242 | hCoV-19/USA/NV-NSPHL-A0196/2020\|EPI_ISL_515450\|202 | ···················· | ·A·················· | ···················· | ···················· | ···················· | ···················· |
| F | 2243 | hCoV-19/USA/NV-NSPHL-A0197/2020\|EPI_ISL_515451\|202 | ···················· | ···················· | ···················· | ···················· | ···················· | ···················· |
| F | 2244 | hCoV-19/USA/NV-NSPHL-A0198/2020\|EPI_ISL_515452\|202 | ···················· | ···················· | ···················· | ···················· | ···················· | ···················· |
| F | 2245 | hCoV-19/USA/NV-NSPHL-A0199/2020\|EPI_ISL_515453\|202 | ···················· | ···················· | ···················· | ···················· | ···················· | ···················· |
| F | 2246 | hCoV-19/USA/NV-NSPHL-A0200/2020\|EPI_ISL_515454\|202 | ···················· | ·A·················· | ···················· | ···················· | ···················· | ···················· |
| F | 2247 | hCoV-19/USA/NV-NSPHL-A0201/2020\|EPI_ISL_515455\|202 | ···················· | ·A·················· | ···················· | ···················· | ···················· | ···················· |
| F | 2248 | hCoV-19/USA/NV-NSPHL-A0202/2020\|EPI_ISL_515456\|202 | ···················· | ···················· | ···················· | ···················· | ···················· | ···················· |
| F | 2249 | hCoV-19/USA/NV-NSPHL-A0203/2020\|EPI_ISL_515457\|202 | ···················· | ···················· | ···················· | ···················· | ···················· | ···················· |
| F | 2250 | hCoV-19/USA/NV-NSPHL-A0205/2020\|EPI_ISL_515459\|202 | ···················· | ···················· | ···················· | ···················· | ···················· | ···················· |
| F | 2251 | hCoV-19/USA/NV-NSPHL-A0206/2020\|EPI_ISL_515460\|202 | ···················· | ···················· | ···················· | ···················· | ···················· | ···················· |
| F | 2252 | hCoV-19/USA/NV-NSPHL-A0208/2020\|EPI_ISL_515461\|202 | ···················· | ···················· | ···················· | ···················· | ···················· | ···················· |
| F | 2253 | hCoV-19/USA/NV-NSPHL-A0210/2020\|EPI_ISL_515462\|202 | ···················· | ···················· | ···················· | ···················· | ···················· | ···················· |
| F | 2254 | hCoV-19/Thailand/Bangkok_3001/2020\|EPI_ISL_515463\| | ···················· | ···················· | ···················· | ···················· | ···················· | ···················· |
| F | 2255 | hCoV-19/Thailand/NIH-3013/2020\|EPI_ISL_515464\|2020 | ···················· | ···················· | ···················· | ···················· | ···················· | ···················· |
| F | 2256 | hCoV-19/Thailand/Bangkok_3049/2020\|EPI_ISL_515465\| | ···················· | ···················· | ···················· | ···················· | ···················· | ···················· |
| F | 2257 | hCoV-19/Canada/AB-Z0002/2020\|EPI_ISL_515466\|2020-0 | ···················· | ···················· | ···················· | ···················· | ···················· | ···················· |
| F | 2258 | hCoV-19/Thailand/Bangkok_3055/2020\|EPI_ISL_515468\| | ···················· | ···················· | ···················· | ···················· | ···················· | ···················· |
| F | 2259 | hCoV-19/Thailand/Nonthaburi_3059/2020\|EPI_ISL_5154 | ···················· | ···················· | ···················· | ···················· | ···················· | ···················· |
| F | 2260 | hCoV-19/Thailand/Nonthaburi_2953/2020\|EPI_ISL_5154 | ···················· | ···················· | ···················· | ···················· | ···················· | ···················· |
| F | 2261 | hCoV-19/Thailand/Bangkok_2848/2020\|EPI_ISL_515472\| | ···················· | ···················· | ···················· | ···················· | ···················· | ···················· |
| F | 2262 | hCoV-19/Thailand/Bangkok_2854/2020\|EPI_ISL_515473\| | ···················· | ···················· | ···················· | ···················· | ···················· | ···················· |
| F | 2263 | hCoV-19/Thailand/Bangkok_2864/2020\|EPI_ISL_515475\| | ···················· | ···················· | ···················· | ···················· | ···················· | ···················· |
| F | 2264 | hCoV-19/Thailand/Bangkok_2866/2020\|EPI_ISL_515476\| | ···················· | ···················· | ···················· | ···················· | ···················· | ···················· |

```
F 2265 hCoV-19/Thailand/Bangkok_2867/2020|EPI_ISL_515477| ···················· ···················· ···················· ···················· ···················· ····················
F 2266 hCoV-19/USA/VA-DCLS-0826/2020|EPI_ISL_516700|2020- ···················· ···················· ···················· ···················· ···················· ····················
F 2267 hCoV-19/USA/VA-DCLS-0828/2020|EPI_ISL_516701|2020- ···················· ···················· ···················· ···················· ···················· ····················
F 2268 hCoV-19/USA/VA-DCLS-0829/2020|EPI_ISL_516702|2020- ···················· ···················· ···················· ···················· ···················· ····················
F 2269 hCoV-19/USA/VA-DCLS-0831/2020|EPI_ISL_516703|2020- ···················· ···················· ···················· ···················· ···················· ····················
F 2270 hCoV-19/USA/VA-DCLS-0835/2020|EPI_ISL_516704|2020- ···················· ···················· ···················· ···················· ···················· ····················
F 2271 hCoV-19/USA/VA-DCLS-0838/2020|EPI_ISL_516705|2020- ···················· ···················· ···················· ···················· ···················· ····················
F 2272 hCoV-19/USA/VA-DCLS-0839/2020|EPI_ISL_516706|2020- ···················· ···················· ···················· ···················· ···················· ····················
F 2273 hCoV-19/USA/VA-DCLS-0840/2020|EPI_ISL_516707|2020- ···················· ···················· ···················· ···················· ···················· ····················
F 2274 hCoV-19/USA/VA-DCLS-0845/2020|EPI_ISL_516710|2020- ···················· ···················· ···················· ···················· ···················· ····················
F 2275 hCoV-19/USA/VA-DCLS-0846/2020|EPI_ISL_516711|2020- ···················· ···················· ···················· ···················· ···················· ····················
F 2276 hCoV-19/USA/VA-DCLS-0848/2020|EPI_ISL_516712|2020- ···················· ···················· ···················· ···················· ···················· ····················
F 2277 hCoV-19/USA/VA-DCLS-0849/2020|EPI_ISL_516713|2020- ···················· ···················· ···················· ···················· ···················· ····················
F 2278 hCoV-19/USA/VA-DCLS-0850/2020|EPI_ISL_516714|2020- ···················· ···················· ···················· ···················· ···················· ····················
F 2279 hCoV-19/USA/VA-DCLS-0851/2020|EPI_ISL_516715|2020- ···················· ···················· ···················· ···················· ···················· ····················
F 2280 hCoV-19/USA/SC-20-0946-T07/2020|EPI_ISL_516749|202 ···················· ···················· ···················· ···················· ···················· ····················
F 2281 hCoV-19/USA/SC-20-0946-T10/2020|EPI_ISL_516750|202 ···················· ···················· ···················· ···················· ···················· ····················
F 2283 hCoV-19/USA/SC-20-1273-T07/2020|EPI_ISL_516753|202 ···················· ···················· ···················· ···················· ···················· ····················
F 2284 hCoV-19/USA/SC-20-1385-T10/2020|EPI_ISL_516755|202 ···················· ···················· ···················· ···················· ···················· ····················
F 2285 hCoV-19/USA/SC-20-1389-T14/2020|EPI_ISL_516756|202 ···················· ···················· ···················· ···················· ···················· ····················
F 2286 hCoV-19/USA/SC-20-1392-T14/2020|EPI_ISL_516757|202 ···················· ···················· ···················· ···················· ···················· ····················
F 2287 hCoV-19/USA/SC-20-1398-T14/2020|EPI_ISL_516758|202 ···················· ···················· ···················· ···················· ···················· ····················
F 2288 hCoV-19/USA/SC-20-1413-T10/2020|EPI_ISL_516762|202 ···················· ···················· ···················· ···················· ···················· ····················
F 2289 hCoV-19/USA/SC-20-1415-T10/2020|EPI_ISL_516763|202 ···················· ···················· ···················· ···················· ···················· ····················
F 2290 hCoV-19/USA/SC-20-1418-T07/2020|EPI_ISL_516764|202 ···················· ···················· ···················· ···················· ···················· ····················
F 2291 hCoV-19/USA/SC-20-1419-T14/2020|EPI_ISL_516765|202 ···················· ···················· ···················· ···················· ···················· ····················
F 2292 hCoV-19/USA/SC-20-1448-T14/2020|EPI_ISL_516766|202 ···················· ···················· ···················· ···················· ···················· ····················
F 2293 hCoV-19/USA/SC-20-1505-T03/2020|EPI_ISL_516767|202 ···················· ···················· ···················· ···················· ···················· ····················
F 2296 hCoV-19/USA/SC-20-1588-T07/2020|EPI_ISL_516771|202 ···················· ···················· ···················· ···················· ···················· ····················
F 2297 hCoV-19/USA/SC-20-1597-T14/2020|EPI_ISL_516772|202 ···················· ···················· ···················· ···················· ···················· ····················
F 2298 hCoV-19/USA/SC-20-1601-T03/2020|EPI_ISL_516773|202 ···················· ···················· ···················· ···················· ···················· ····················
F 2299 hCoV-19/USA/SC-20-1609-T07/2020|EPI_ISL_516774|202 ···················· ···················· ···················· ···················· ···················· ····················
F 2300 hCoV-19/USA/SC-20-1615-T14/2020|EPI_ISL_516775|202 ···················· ···················· ···················· ···················· ···················· ····················
F 2301 hCoV-19/USA/SC-20-1633-T07/2020|EPI_ISL_516776|202 ···················· ·········A·········· ···················· ···················· ···················· ····················
F 2302 hCoV-19/USA/SC-20-1803-T07/2020|EPI_ISL_516777|202 ···················· ···················· ···················· ···················· ···················· ····················
F 2303 hCoV-19/USA/SC-20-1820-T07/2020|EPI_ISL_516778|202 ···················· ···················· ···················· ···················· ···················· ····················
F 2305 hCoV-19/USA/WA-S2493/2020|EPI_ISL_497125|2020-06-1 ···················· ···················· ···················· ···················· ···················· ····················
F 2306 hCoV-19/USA/WA-S2494/2020|EPI_ISL_497126|2020-06-1 ···················· ···················· ···················· ···················· ···················· ····················
F 2307 hCoV-19/USA/WA-S2495/2020|EPI_ISL_497127|2020-06-1 ···················· ···················· ···················· ···················· ···················· ····················
F 2308 hCoV-19/USA/WA-S2496/2020|EPI_ISL_497128|2020-06-1 ···················· ···················· ···················· ···················· ···················· ····················
F 2309 hCoV-19/USA/WA-S2497/2020|EPI_ISL_497129|2020-06-1 ···················· ···················· ···················· ···················· ···················· ····················
F 2310 hCoV-19/USA/WA-S2504/2020|EPI_ISL_497136|2020-06-1 ···················· ···················· ···················· ···················· ···················· ····················
F 2311 hCoV-19/USA/WA-S2505/2020|EPI_ISL_497137|2020-06-1 ···················· ···················· ···················· ···················· ···················· ····················
F 2312 hCoV-19/USA/WA-S2506/2020|EPI_ISL_497138|2020-06-1 ···················· ···················· ···················· ···················· ···················· ····················
F 2313 hCoV-19/USA/WA-S2507/2020|EPI_ISL_497139|2020-06-1 ···················· ···················· ···················· ···················· ···················· ····················
F 2314 hCoV-19/USA/WA-S2508/2020|EPI_ISL_497140|2020-06-1 ···················· ···················· ···················· ···················· ···················· ····················
F 2315 hCoV-19/USA/WA-S2509/2020|EPI_ISL_497141|2020-06-1 ···················· ···················· ···················· ···················· ···················· ····················
F 2316 hCoV-19/USA/WA-S2510/2020|EPI_ISL_497142|2020-06-1 ···················· ···················· ···················· ···················· ···················· ····················
F 2317 hCoV-19/USA/WA-S2511/2020|EPI_ISL_497143|2020-06-1 ···················· ···················· ···················· ···················· ···················· ····················
F 2318 hCoV-19/USA/WA-S2512/2020|EPI_ISL_497144|2020-06-1 ···················· ···················· ···················· ···················· ···················· ····················
F 2319 hCoV-19/USA/WA-S2513/2020|EPI_ISL_497145|2020-06-1 ···················· ···················· ···················· ···················· ···················· ····················
F 2320 hCoV-19/USA/WA-S2514/2020|EPI_ISL_497146|2020-06-1 ···················· ···················· ···················· ···················· ···················· ····················
F 2321 hCoV-19/USA/WA-S2537/2020|EPI_ISL_497169|2020-06-1 ···················· ···················· ···················· ···················· ···················· ····················
F 2322 hCoV-19/USA/WA-S2538/2020|EPI_ISL_497170|2020-06-1 ···················· ···················· ···················· ···················· ···················· ····················
F 2323 hCoV-19/USA/WA-S2539/2020|EPI_ISL_497171|2020-06-1 ···················· ···················· ···················· ···················· ···················· ····················
F 2324 hCoV-19/USA/WA-S2540/2020|EPI_ISL_497172|2020-06-1 ···················· ···················· ···················· ···················· ···················· ····················
F 2325 hCoV-19/USA/WA-S2541/2020|EPI_ISL_497173|2020-06-1 ···················· ···················· ···················· ···················· ···················· ····················
F 2326 hCoV-19/USA/WA-S2542/2020|EPI_ISL_497174|2020-06-1 ···················· ···················· ···················· ···················· ···················· ····················
F 2327 hCoV-19/USA/WA-S2543/2020|EPI_ISL_497175|2020-06-1 ···················· ···················· ···················· ···················· ···················· ····················
F 2328 hCoV-19/USA/WA-S2544/2020|EPI_ISL_497176|2020-06-1 ···················· ···················· ···················· ···················· ···················· ····················
F 2329 hCoV-19/USA/WA-S2545/2020|EPI_ISL_497177|2020-06-1 ···················· ···················· ···················· ···················· ···················· ····················
F 2330 hCoV-19/USA/WA-S2546/2020|EPI_ISL_497178|2020-06-1 ···················· ···················· ···················· ···················· ···················· ····················
F 2331 hCoV-19/USA/WA-S2547/2020|EPI_ISL_497179|2020-06-1 ···················· ···················· ···················· ···················· ···················· ····················
F 2332 hCoV-19/USA/WA-S2548/2020|EPI_ISL_497180|2020-06-1 ···················· ···················· ···················· ···················· ···················· ····················
F 2333 hCoV-19/USA/WA-S2549/2020|EPI_ISL_497181|2020-06-1 ···················· ···················· ···················· ···················· ···················· ····················
F 2334 hCoV-19/USA/WA-S2550/2020|EPI_ISL_497182|2020-06-1 ···················· ···················· ···················· ···················· ···················· ····················
F 2335 hCoV-19/USA/WA-S2551/2020|EPI_ISL_497183|2020-06-1 ···················· ···················· ···················· ···················· ···················· ····················
F 2336 hCoV-19/USA/WA-S2552/2020|EPI_ISL_497184|2020-06-1 ···················· ···················· ···················· ···················· ···················· ····················
F 2337 hCoV-19/USA/WA-S2553/2020|EPI_ISL_497185|2020-06-1 ···················· ···················· ···················· ···················· ···················· ····················
F 2338 hCoV-19/USA/WA-S2554/2020|EPI_ISL_497186|2020-06-1 ···················· ···················· ···················· ···················· ···················· ····················
F 2339 hCoV-19/USA/WA-S2555/2020|EPI_ISL_497187|2020-06-1 ···················· ···················· ···················· ···················· ···················· ····················
F 2340 hCoV-19/USA/WA-S2557/2020|EPI_ISL_497189|2020-06-1 ···················· ···················· ···················· ···················· ···················· ····················
F 2341 hCoV-19/USA/WA-S2558/2020|EPI_ISL_497190|2020-06-1 ···················· ···················· ···················· ···················· ···················· ····················
F 2342 hCoV-19/USA/WA-S2559/2020|EPI_ISL_497191|2020-06-1 ···················· ···················· ···················· ···················· ···················· ····················
F 2343 hCoV-19/USA/WA-S2560/2020|EPI_ISL_497192|2020-06-1 ···················· ···················· ···················· ···················· ···················· ····················
F 2344 hCoV-19/USA/WA-S2561/2020|EPI_ISL_497193|2020-06-1 ···················· ···················· ···················· ···················· ···················· ····················
F 2345 hCoV-19/USA/WA-S2562/2020|EPI_ISL_497194|2020-06-1 ···················· ···················· ···················· ···················· ···················· ····················
F 2346 hCoV-19/USA/WA-S2563/2020|EPI_ISL_497195|2020-06-1 ···················· ···················· ···················· ···················· ···················· ····················
F 2347 hCoV-19/USA/WA-S2564/2020|EPI_ISL_497196|2020-06-1 ···················· ···················· ···················· ···················· ···················· ····················
F 2348 hCoV-19/USA/WA-S2565/2020|EPI_ISL_497197|2020-06-1 ···················· ···················· ···················· ···················· ···················· ····················
```

F 2349 hCoV-19/USA/WA-S2566/2020|EPI_ISL_497198|2020-06-1 ···················· ···················· ····················· ···················· ···················· ····················
F 2350 hCoV-19/USA/WA-S2567/2020|EPI_ISL_497199|2020-06-1 ···················· ···················· ····················· ···················· ···················· ····················
F 2351 hCoV-19/Bangladesh/BCSIR-NILMRC-285/2020|EPI_ISL_4 ···················· ···················· ····················· ···················· ···················· ····················
F 2352 hCoV-19/Australia/ACT0001/2020|EPI_ISL_498468|2020 ···················· ···················· ····················· ···················· ···················· ····················
F 2353 hCoV-19/Australia/ACT0005/2020|EPI_ISL_498472|2020 ···················· ···················· ····················· ···················· ···················· ····················
F 2354 hCoV-19/Australia/ACT0006/2020|EPI_ISL_498473|2020 ···················· ···················· ····················· ···················· ···················· ····················
F 2355 hCoV-19/Australia/ACT0010/2020|EPI_ISL_498477|2020 ···················· ···················· ····················· ···················· ···················· ····················
F 2356 hCoV-19/England/NORT-29C650/2020|EPI_ISL_499787|20 ···················· ···················· ····················· ···················· ···················· ····················
F 2357 hCoV-19/England/NORT-29C7E4/2020|EPI_ISL_499788|20 ···················· ···················· ····················· ···················· ···················· ····················
F 2358 hCoV-19/England/NORT-29CD00/2020|EPI_ISL_499789|20 ···················· ···················· ····················· ···················· ···················· ····················
F 2359 hCoV-19/England/NORT-29D17C/2020|EPI_ISL_499798|20 ···················· ···················· ····················· ···················· ···················· ····················
F 2360 hCoV-19/England/NORT-29D1B8/2020|EPI_ISL_499799|20 ···················· ···················· ····················· ···················· ···················· ····················
F 2361 hCoV-19/Australia/SAP437/2020|EPI_ISL_508128|2020- ···················· ···················· ····················· ···················· ···················· ····················
F 2362 hCoV-19/Australia/SAP438/2020|EPI_ISL_508129|2020- ···················· ···················· ····················· ···················· ···················· ····················
F 2363 hCoV-19/Australia/SAP439/2020|EPI_ISL_508130|2020- ···················· ···················· ····················· ···················· ···················· ····················
F 2364 hCoV-19/Australia/SAP440/2020|EPI_ISL_508131|2020- ···················· ···················· ····················· ···················· ···················· ····················
F 2365 hCoV-19/Australia/SAP441/2020|EPI_ISL_508132|2020- ···················· ···················· ····················· ···················· ···················· ····················
F 2366 hCoV-19/Australia/SAP442/2020|EPI_ISL_508133|2020- ···················· ···········A········ ····················· ···················· ···················· ····················
F 2367 hCoV-19/Australia/SAP443/2020|EPI_ISL_508134|2020- ···················· ···················· ····················· ···················· ···················· ····················
F 2368 hCoV-19/Australia/SAP444/2020|EPI_ISL_508135|2020- ···················· ·············A······ ····················· ···················· ···················· ····················
F 2369 hCoV-19/Australia/SAP445/2020|EPI_ISL_508136|2020- ···················· ···················· ····················· ···················· ···················· ····················
F 2370 hCoV-19/Australia/SAP446/2020|EPI_ISL_508137|2020- ···················· ···················· ····················· ···················· ···················· ····················
F 2371 hCoV-19/Australia/SAP447/2020|EPI_ISL_508138|2020- ···················· ···················· ····················· ···················· ···················· ····················
F 2372 hCoV-19/Australia/SAP448/2020|EPI_ISL_508139|2020- ···················· ···················· ····················· ···················· ···················· ····················
F 2373 hCoV-19/Australia/SAP450/2020|EPI_ISL_508140|2020- ···················· ···················· ····················· ···················· ···················· ····················
F 2374 hCoV-19/Australia/SAP451/2020|EPI_ISL_508141|2020- ···················· ···················· ····················· ···················· ···················· ····················
F 2375 hCoV-19/Australia/SAP452/2020|EPI_ISL_508142|2020- ···················· ···················· ····················· ···················· ···················· ····················
F 2377 hCoV-19/India/UT-AR1/2020|EPI_ISL_508156|2020-06-0 ···················· ···················· ····················· ···················· ···················· ····················
F 2378 hCoV-19/India/UT-AR10/2020|EPI_ISL_508157|2020-05- ···················· ···················· ····················· ···················· ···················· ····················
F 2379 hCoV-19/India/UT-AR11/2020|EPI_ISL_508158|2020-05- ···················· ···················· ····················· ···················· ···················· ····················
F 2380 hCoV-19/India/UT-AR12/2020|EPI_ISL_508159|2020-05- ···················· ···················· ····················· ···················· ···················· ····················
F 2381 hCoV-19/India/UT-AR13/2020|EPI_ISL_508160|2020-06- ···················· ·A·················· ····················· ···················· ···················· ···A················
F 2382 hCoV-19/India/UT-AR14/2020|EPI_ISL_508161|2020-06- ···················· ·A·················· ····················· ···················· ···················· ···A················
F 2383 hCoV-19/India/UT-AR15/2020|EPI_ISL_508162|2020-05- ···················· ···················· ····················· ···················· ···················· ····················
F 2384 hCoV-19/India/UT-AR16/2020|EPI_ISL_508163|2020-05- ···················· ···················· ····················· ···················· ···················· ····················
F 2385 hCoV-19/India/UT-AR17/2020|EPI_ISL_508164|2020-05- ···················· ·A·················· ····················· ···················· ···················· ···A················
F 2386 hCoV-19/India/UT-AR18/2020|EPI_ISL_508165|2020-05- ···················· ···················· ····················· ···················· ···················· ····················
F 2387 hCoV-19/India/UT-AR19/2020|EPI_ISL_508166|2020-05- ···················· ···················· ····················· ···················· ···················· ····················
F 2388 hCoV-19/India/UT-AR2/2020|EPI_ISL_508167|2020-05-3 ···················· ···················· ····················· ···················· ···················· ····················
F 2389 hCoV-19/India/UP-AR20/2020|EPI_ISL_508168|2020-06- ···················· ·A·················· ····················· ···················· ···················· ···A················
F 2390 hCoV-19/India/UT-AR21/2020|EPI_ISL_508169|2020-05- ···················· ···················· ····················· ···················· ···················· ····················
F 2391 hCoV-19/India/KA-IB22/2020|EPI_ISL_508300|2020-05- ···················· ···················· ····················· ···················· ···················· ····················
F 2392 hCoV-19/India/KA-IB23/2020|EPI_ISL_508301|2020-06- ···················· ········A··········· ····················· ···················· ···················· ····················
F 2393 hCoV-19/India/KA-IB24/2020|EPI_ISL_508302|2020-06- ···················· ···················· ····················· ···················· ···················· ····················
F 2394 hCoV-19/India/KA-IB25/2020|EPI_ISL_508303|2020-06- ···················· ···················· ····················· ···················· ···················· ····················
F 2395 hCoV-19/India/KA-IB28/2020|EPI_ISL_508306|2020-06- ···················· ···················· ····················· ···················· ···················· ····················
F 2396 hCoV-19/India/KA-IB3/2020|EPI_ISL_508308|2020-06-1 ···················· ···················· ····················· ···················· ···················· ····················
F 2397 hCoV-19/India/KA-IB30/2020|EPI_ISL_508309|2020-05- ···················· ···················· ····················· ···················· ···················· ····················
F 2398 hCoV-19/England/PRIN-254BCA0/2020|EPI_ISL_509440|2 ···················· ···················· ····················· ···················· ···················· ····················
F 2399 hCoV-19/England/PRIN-254BB49/2020|EPI_ISL_509442|2 ···················· ···················· ····················· ···················· ···················· ····················
F 2400 hCoV-19/England/PRIN-254BD34/2020|EPI_ISL_509443|2 ···················· ···················· ····················· ···················· ···················· ····················
F 2401 hCoV-19/USA/MD-MDH-0040/2020|EPI_ISL_509444|2020-0 ···················· ···················· ····················· ···················· ···················· ····················
F 2402 hCoV-19/USA/MD-MDH-0041/2020|EPI_ISL_509445|2020-0 ···················· ···················· ····················· ···················· ···················· ····················
F 2403 hCoV-19/USA/MD-MDH-0042/2020|EPI_ISL_509446|2020-0 ···················· ···················· ····················· ···················· ···················· ····················
F 2404 hCoV-19/USA/MD-MDH-0043/2020|EPI_ISL_509447|2020-0 ···················· ···················· ····················· ···················· ···················· ····················
F 2405 hCoV-19/USA/MD-MDH-0044/2020|EPI_ISL_509448|2020-0 ···················· ···················· ····················· ···················· ···················· ····················
F 2406 hCoV-19/USA/MD-MDH-0045/2020|EPI_ISL_509449|2020-0 ···················· ···················· ····················· ···················· ···················· ····················
F 2407 hCoV-19/USA/MD-MDH-0055/2020|EPI_ISL_509459|2020-0 ···················· ···················· ····················· ···················· ···················· ····················
F 2408 hCoV-19/USA/MD-MDH-0056/2020|EPI_ISL_509460|2020-0 ···················· ···················· ····················· ···················· ···················· ····················
F 2409 hCoV-19/USA/MD-MDH-0057/2020|EPI_ISL_509461|2020-0 ···················· ···················· ····················· ···················· ···················· ····················
F 2410 hCoV-19/USA/MD-MDH-0058/2020|EPI_ISL_509462|2020-0 ···················· ···················· ····················· ···················· ···················· ····················
F 2411 hCoV-19/USA/MD-MDH-0059/2020|EPI_ISL_509463|2020-0 ···················· ···················· ····················· ···················· ···················· ····················
F 2412 hCoV-19/USA/MD-MDH-0060/2020|EPI_ISL_509464|2020-0 ···················· ···················· ····················· ···················· ···················· ····················
F 2413 hCoV-19/USA/MD-MDH-0061/2020|EPI_ISL_509465|2020-0 ···················· ···················· ····················· ···················· ···················· ····················
F 2414 hCoV-19/USA/MD-MDH-0062/2020|EPI_ISL_509466|2020-0 ···················· ···················· ····················· ···················· ···················· ····················
F 2415 hCoV-19/USA/MD-MDH-0063/2020|EPI_ISL_509467|2020-0 ···················· ···················· ····················· ···················· ···················· ····················
F 2416 hCoV-19/USA/MD-MDH-0064/2020|EPI_ISL_509468|2020-0 ···················· ···················· ····················· ···················· ···················· ····················
F 2417 hCoV-19/USA/MD-MDH-0065/2020|EPI_ISL_509469|2020-0 ···················· ···················· ····················· ···················· ···················· ····················
F 2418 hCoV-19/USA/MD-MDH-0066/2020|EPI_ISL_509470|2020-0 ···················· ···················· ····················· ···················· ···················· ····················
F 2419 hCoV-19/USA/MD-MDH-0067/2020|EPI_ISL_509471|2020-0 ···················· ···················· ····················· ···················· ···················· ····················
F 2420 hCoV-19/USA/MD-MDH-0068/2020|EPI_ISL_509472|2020-0 ···················· ···················· ····················· ···················· ···················· ····················
F 2421 hCoV-19/USA/MD-MDH-0069/2020|EPI_ISL_509473|2020-0 ···················· ···················· ····················· ···················· ···················· ····················
F 2422 hCoV-19/USA/MD-MDH-0070/2020|EPI_ISL_509474|2020-0 ···················· ···················· ····················· ···················· ···················· ····················
F 2423 hCoV-19/USA/MD-MDH-0071/2020|EPI_ISL_509475|2020-0 ···················· ···················· ····················· ···················· ···················· ····················
F 2424 hCoV-19/USA/MD-MDH-0072/2020|EPI_ISL_509476|2020-0 ···················· ···················· ····················· ···················· ···················· ····················
F 2425 hCoV-19/USA/MD-MDH-0073/2020|EPI_ISL_509477|2020-0 ···················· ···················· ····················· ···················· ···················· ····················
F 2426 hCoV-19/USA/MD-MDH-0074/2020|EPI_ISL_509478|2020-0 ···················· ···················· ····················· ···················· ···················· ····················
F 2427 hCoV-19/USA/MD-MDH-0075/2020|EPI_ISL_509479|2020-0 ···················· ···················· ····················· ···················· ···················· ····················
F 2428 hCoV-19/USA/MD-MDH-0076/2020|EPI_ISL_509480|2020-0 ···················· ···················· ····················· ···················· ···················· ····················
F 2429 hCoV-19/USA/MD-MDH-0077/2020|EPI_ISL_509481|2020-0 ···················· ···················· ····················· ···················· ···················· ····················

| | | | | | | | | |
|---|---|---|---|---|---|---|---|---|
| F | 2430 | hCoV-19/USA/MD-MDH-0078/2020\|EPI_ISL_509482\|2020-0 | ···················· | ···················· | ····················· | ···················· | ···················· | ···················· |
| F | 2431 | hCoV-19/USA/MD-MDH-0079/2020\|EPI_ISL_509483\|2020-0 | ···················· | ···················· | ····················· | ···················· | ···················· | ···················· |
| F | 2432 | hCoV-19/USA/MD-MDH-0080/2020\|EPI_ISL_509484\|2020-0 | ···················· | ···················· | ····················· | ···················· | ···················· | ···················· |
| F | 2433 | hCoV-19/USA/MD-MDH-0081/2020\|EPI_ISL_509485\|2020-0 | ···················· | ···················· | ····················· | ···················· | ···················· | ···················· |
| F | 2434 | hCoV-19/USA/MD-MDH-0082/2020\|EPI_ISL_509486\|2020-0 | ···················· | ···················· | ····················· | ···················· | ···················· | ···················· |
| F | 2435 | hCoV-19/USA/MD-MDH-0083/2020\|EPI_ISL_509487\|2020-0 | ···················· | ···················· | ····················· | ···················· | ···················· | ···················· |
| F | 2436 | hCoV-19/USA/MD-MDH-0084/2020\|EPI_ISL_509488\|2020-0 | ···················· | ···················· | ····················· | ···················· | ···················· | ···················· |
| F | 2437 | hCoV-19/USA/MD-MDH-0085/2020\|EPI_ISL_509489\|2020-0 | ···················· | ···················· | ····················· | ···················· | ···················· | ···················· |
| F | 2438 | hCoV-19/USA/MD-MDH-0086/2020\|EPI_ISL_509490\|2020-0 | ···················· | ···················· | ····················· | ···················· | ···················· | ···················· |
| F | 2439 | hCoV-19/USA/MD-MDH-0087/2020\|EPI_ISL_509491\|2020-0 | ···················· | ···················· | ····················· | ···················· | ···················· | ···················· |
| F | 2440 | hCoV-19/Australia/NSW2029/2020\|EPI_ISL_509492\|2020 | ···················· | ·············A······ | ····················· | ···················· | ···················· | ···················· |
| F | 2441 | hCoV-19/Australia/NSW2034/2020\|EPI_ISL_509493\|2020 | ···················· | ·············A······ | ····················· | ···················· | ···················· | ···················· |
| F | 2442 | hCoV-19/Australia/NSW2051/2020\|EPI_ISL_509494\|2020 | ···················· | ···················· | ····················· | ···················· | ···················· | ···················· |
| F | 2443 | hCoV-19/Australia/NSW2055/2020\|EPI_ISL_509495\|2020 | ···················· | ···················· | ····················· | ···················· | ···················· | ···················· |
| F | 2444 | hCoV-19/Australia/NSW2063/2020\|EPI_ISL_509496\|2020 | ···················· | ···················· | ····················· | ···················· | ···················· | ···················· |
| F | 2445 | hCoV-19/Australia/NSW2071/2020\|EPI_ISL_509497\|2020 | ···················· | ···················· | ····················· | ···················· | ···················· | ···················· |
| F | 2446 | hCoV-19/Australia/NSW2120/2020\|EPI_ISL_509498\|2020 | ···················· | ···················· | ····················· | ···················· | ···················· | ···················· |
| F | 2447 | hCoV-19/Australia/NSW2125/2020\|EPI_ISL_509499\|2020 | ···················· | ···················· | ····················· | ···················· | ···················· | ···················· |
| F | 2448 | hCoV-19/Spain/PV-IBV-001537/2020\|EPI_ISL_509616\|20 | ···················· | ···················· | ····················· | ···················· | ···················· | ···················· |
| F | 2449 | hCoV-19/Spain/PV-IBV-001538/2020\|EPI_ISL_509617\|20 | ···················· | ···················· | ····················· | ···················· | ···················· | ···················· |
| F | 2450 | hCoV-19/Spain/PV-IBV-001542/2020\|EPI_ISL_509618\|20 | ···················· | ···················· | ····················· | ···················· | ···················· | ···················· |
| F | 2451 | hCoV-19/Spain/AN-IBV-001980/2020\|EPI_ISL_509619\|20 | ···················· | ···················· | ····················· | ···················· | ···················· | ···················· |
| F | 2452 | hCoV-19/Spain/AN-IBV-002020/2020\|EPI_ISL_509630\|20 | ···················· | ···················· | ····················· | ···················· | ···················· | ···················· |
| F | 2453 | hCoV-19/Spain/AN-IBV-002023/2020\|EPI_ISL_509631\|20 | ···················· | ···················· | ····················· | ···················· | ···················· | ···················· |
| F | 2454 | hCoV-19/Spain/AN-IBV-002024/2020\|EPI_ISL_509632\|20 | ···················· | ···················· | ····················· | ···················· | ···················· | ···················· |
| F | 2455 | hCoV-19/Spain/PV-IBV-005137/2020\|EPI_ISL_509633\|20 | ···················· | ···················· | ····················· | ···················· | ···················· | ···················· |
| F | 2456 | hCoV-19/Spain/PV-IBV-005139/2020\|EPI_ISL_509634\|20 | ···················· | ···················· | ····················· | ···················· | ···················· | ···················· |
| F | 2457 | hCoV-19/Spain/PV-IBV-005149/2020\|EPI_ISL_509639\|20 | ···················· | ···················· | ····················· | ···················· | ···················· | ···················· |
| F | 2458 | hCoV-19/Spain/AN-IBV-006080/2020\|EPI_ISL_510424\|20 | ···················· | ···················· | ····················· | ···················· | ···················· | ···················· |
| F | 2468 | hCoV-19/Spain/AN-IBV-006116/2020\|EPI_ISL_510445\|20 | ···················· | ···················· | ····················· | ···················· | ···················· | ···················· |
| F | 2475 | hCoV-19/Spain/AN-IBV-006123/2020\|EPI_ISL_510452\|20 | ···················· | ···················· | ····················· | ···················· | ···················· | ···················· |
| F | 2478 | hCoV-19/Spain/AN-IBV-006130/2020\|EPI_ISL_510456\|20 | ···················· | ···················· | ····················· | ···················· | ···················· | ···················· |
| F | 2485 | hCoV-19/Spain/PV-IBV-004180/2020\|EPI_ISL_510467\|20 | ···················· | ···················· | ····················· | ···················· | ···················· | ···················· |
| F | 2486 | hCoV-19/Spain/PV-IBV-004181/2020\|EPI_ISL_510468\|20 | ···················· | ···················· | ····················· | ···················· | ···················· | ···················· |
| F | 2488 | hCoV-19/Spain/PV-IBV-004183/2020\|EPI_ISL_510470\|20 | ···················· | ···················· | ····················· | ···················· | ···················· | ···················· |
| F | 2489 | hCoV-19/Spain/PV-IBV-004184/2020\|EPI_ISL_510471\|20 | ···················· | ···················· | ····················· | ···················· | ···················· | ···················· |
| F | 2491 | hCoV-19/Spain/PV-IBV-004187/2020\|EPI_ISL_510474\|20 | ···················· | ···················· | ····················· | ···················· | ·NN················· | ···················· |
| F | 2498 | hCoV-19/Spain/PV-IBV-004195/2020\|EPI_ISL_510481\|20 | ···················· | ···················· | ····················· | ···················· | ··NN················ | N··················· |
| F | 2507 | hCoV-19/South Korea/KCDC2262/2020\|EPI_ISL_510600\|2 | ···················· | ···················· | ····················· | ···················· | ···················· | ···················· |
| F | 2508 | hCoV-19/South Korea/KCDC2263/2020\|EPI_ISL_510601\|2 | ···················· | ···················· | ····················· | ···················· | ···················· | ···················· |
| F | 2509 | hCoV-19/South Korea/KCDC2264/2020\|EPI_ISL_510602\|2 | ···················· | ···················· | ····················· | ···················· | ···················· | ···················· |
| F | 2510 | hCoV-19/South Korea/KCDC2265/2020\|EPI_ISL_510603\|2 | ···················· | ···················· | ····················· | ···················· | ···················· | ···················· |
| F | 2511 | hCoV-19/South Korea/KCDC2270/2020\|EPI_ISL_510608\|2 | ···················· | ···················· | ····················· | ···················· | ···················· | ···················· |
| F | 2512 | hCoV-19/South Korea/KCDC2271/2020\|EPI_ISL_510609\|2 | ···················· | ···················· | ····················· | ···················· | ···················· | ···················· |
| F | 2513 | hCoV-19/South Korea/KCDC2273/2020\|EPI_ISL_510610\|2 | ···················· | ···················· | ····················· | ···················· | ···················· | ···················· |
| F | 2514 | hCoV-19/Portugal/PT1212/2020\|EPI_ISL_511755\|2020-0 | ···················· | ···················· | ····················· | ···················· | ···················· | ···················· |
| F | 2515 | hCoV-19/Portugal/PT1213/2020\|EPI_ISL_511756\|2020-0 | ···················· | ···················· | ····················· | ···················· | ···················· | ···················· |
| F | 2516 | hCoV-19/Portugal/PT1214/2020\|EPI_ISL_511757\|2020-0 | ···················· | ···················· | ····················· | ···················· | ···················· | ···················· |
| F | 2517 | hCoV-19/Portugal/PT1216/2020\|EPI_ISL_511759\|2020-0 | ···················· | ···················· | ····················· | ···················· | ···················· | ···················· |
| F | 2518 | hCoV-19/Portugal/PT1223/2020\|EPI_ISL_511766\|2020-0 | ···················· | ···················· | ····················· | ···················· | ···················· | ···················· |
| F | 2519 | hCoV-19/Portugal/PT1224/2020\|EPI_ISL_511767\|2020-0 | ···················· | ···················· | ····················· | ···················· | ···················· | ···················· |
| F | 2520 | hCoV-19/Portugal/PT1225/2020\|EPI_ISL_511768\|2020-0 | ···················· | ···················· | ····················· | ···················· | ···················· | ···················· |
| F | 2521 | hCoV-19/Portugal/PT1226/2020\|EPI_ISL_511769\|2020-0 | ···················· | ···················· | ····················· | ···················· | ···················· | ···················· |
| F | 2522 | hCoV-19/Portugal/PT1227/2020\|EPI_ISL_511770\|2020-0 | ···················· | ···················· | ····················· | ···················· | ···················· | ···················· |
| F | 2523 | hCoV-19/Portugal/PT1228/2020\|EPI_ISL_511771\|2020-0 | ···················· | ···················· | ····················· | ···················· | ···················· | ···················· |
| F | 2524 | hCoV-19/India/WB-IK3/2020\|EPI_ISL_511900\|2020-03-3 | ···················· | ···················· | ····················· | ···················· | ···················· | ···················· |
| F | 2525 | hCoV-19/India/WB-IK8/2020\|EPI_ISL_511901\|2020-04-0 | ···················· | ···················· | ····················· | ···················· | ···················· | ···················· |
| F | 2526 | hCoV-19/India/WB-IK52/2020\|EPI_ISL_511902\|2020-05- | ···················· | ···················· | ····················· | ···················· | ···················· | ···················· |
| F | 2527 | hCoV-19/India/WB-IK62/2020\|EPI_ISL_511903\|2020-04- | ···················· | ···················· | ····················· | ···················· | ···A················ | ···················· |
| F | 2528 | hCoV-19/India/WB-IK70/2020\|EPI_ISL_511904\|2020-05- | ···················· | ···················· | ····················· | ···················· | ···················· | ···················· |
| F | 2529 | hCoV-19/India/WB-IK81/2020\|EPI_ISL_511905\|2020-05- | ···················· | ···················· | ····················· | ···················· | ···················· | ···················· |
| F | 2530 | hCoV-19/India/WB-IK89/2020\|EPI_ISL_511906\|2020-05- | ···················· | ···················· | ····················· | ···················· | ···A················ | ···················· |
| F | 2531 | hCoV-19/India/WB-IK95/2020\|EPI_ISL_511907\|2020-05- | ···················· | ···················· | ····················· | ···················· | ···················· | ···················· |
| F | 2532 | hCoV-19/India/UT-AR26/2020\|EPI_ISL_511908\|2020-05- | ···················· | ···················· | ····················· | ···················· | ···················· | ···················· |
| F | 2533 | hCoV-19/India/UT-AR35/2020\|EPI_ISL_511909\|2020-05- | ···················· | ···················· | ····················· | ···················· | ···················· | ···················· |
| F | 2534 | hCoV-19/India/UT-AR37/2020\|EPI_ISL_511910\|2020-05- | ···················· | ···················· | ····················· | ···················· | ···················· | ···················· |
| F | 2535 | hCoV-19/India/UT-AR40/2020\|EPI_ISL_511912\|2020-05- | ···················· | ···················· | ····················· | ···················· | ···················· | ···················· |
| F | 2536 | hCoV-19/India/UT-AR57/2020\|EPI_ISL_511921\|2020-06- | ···················· | ···················· | ····················· | ···················· | ···················· | ···················· |
| F | 2537 | hCoV-19/India/UT-AR64/2020\|EPI_ISL_511922\|2020-06- | ···················· | ·A·················· | ····················· | ···················· | ···················· | ···A················ |
| F | 2538 | hCoV-19/India/MH-MW22/2020\|EPI_ISL_511924\|2020-05- | ···················· | ···················· | ····················· | ···················· | ···················· | ···················· |
| F | 2539 | hCoV-19/India/MH-MW35/2020\|EPI_ISL_511926\|2020-06- | ···················· | ·A·················· | ····················· | ···················· | ···················· | ···A················ |
| F | 2540 | hCoV-19/India/MH-MW36/2020\|EPI_ISL_511927\|2020-06- | ···················· | ·A·················· | ····················· | ···················· | ···················· | ···A················ |
| F | 2541 | hCoV-19/India/HR-MW39/2020\|EPI_ISL_511928\|2020-06- | ···················· | ···················· | ····················· | ···················· | ···················· | ···················· |
| F | 2542 | hCoV-19/India/MH-MW40/2020\|EPI_ISL_511929\|2020-06- | ···················· | ···················· | ····················· | ···················· | ···················· | ···················· |
| F | 2543 | hCoV-19/India/MH-GA18/2020\|EPI_ISL_511930\|2020-06- | ···················· | ···················· | ····················· | ···················· | ···················· | ···················· |
| F | 2544 | hCoV-19/India/MH-GA30/2020\|EPI_ISL_511931\|2020-06- | ···················· | ···················· | ····················· | ···················· | ···················· | ···················· |
| F | 2545 | hCoV-19/India/MH-GA44/2020\|EPI_ISL_511932\|2020-06- | ···················· | ···················· | ····················· | ···················· | ···················· | ···················· |
| F | 2546 | hCoV-19/India/MH-GA58/2020\|EPI_ISL_511933\|2020-06- | ···················· | ···················· | ····················· | ···················· | ···················· | ···················· |
| F | 2547 | hCoV-19/India/MH-GA72/2020\|EPI_ISL_511934\|2020-06- | ···················· | ···················· | ····················· | ···················· | ···················· | ···················· |
| F | 2548 | hCoV-19/India/MH-GA95/2020\|EPI_ISL_511939\|2020-06- | ···················· | ·A·················· | ····················· | ···················· | ···················· | ···A················ |

| | | | | | | | |
|---|---|---|---|---|---|---|---|
| F | 2549 hCoV-19/USA/FL-BPHL-0825/2020\|EPI_ISL_514171\|2020- | .................... | .................... | .................... | .................... | .................... | .................... |
| F | 2550 hCoV-19/USA/FL-BPHL-0830/2020\|EPI_ISL_514176\|2020- | .................... | .................... | .................... | .................... | .................... | .................... |
| F | 2551 hCoV-19/USA/FL-BPHL-0831/2020\|EPI_ISL_514177\|2020- | .................... | .................... | .................... | .................... | .................... | .................... |
| F | 2552 hCoV-19/USA/FL-BPHL-0832/2020\|EPI_ISL_514178\|2020- | .................... | .................... | .................... | .................... | .................... | .................... |
| F | 2553 hCoV-19/USA/FL-BPHL-0834/2020\|EPI_ISL_514180\|2020- | .................... | .................... | .................... | .................... | .................... | .................... |
| F | 2554 hCoV-19/USA/FL-BPHL-0837/2020\|EPI_ISL_514183\|2020- | .................... | .................... | .................... | .................... | .................... | .................... |
| F | 2555 hCoV-19/USA/FL-BPHL-0838/2020\|EPI_ISL_514184\|2020- | .................... | .................... | .................... | .................... | .................... | .................... |
| F | 2556 hCoV-19/USA/FL-BPHL-0840/2020\|EPI_ISL_514186\|2020- | .................... | .................... | .................... | .................... | .................... | .................... |
| F | 2557 hCoV-19/USA/FL-BPHL-0841/2020\|EPI_ISL_514187\|2020- | .................... | .............A...... | .................... | .................... | .................... | .................... |
| F | 2558 hCoV-19/USA/FL-BPHL-0842/2020\|EPI_ISL_514188\|2020- | .................... | .................... | .................... | .................... | .................... | .................... |
| F | 2559 hCoV-19/USA/FL-BPHL-0843/2020\|EPI_ISL_514189\|2020- | .................... | .................... | .................... | .................... | .................... | .................... |
| F | 2560 hCoV-19/USA/FL-BPHL-0851/2020\|EPI_ISL_514197\|2020- | .................... | .................... | .................... | .................... | .................... | .................... |
| F | 2561 hCoV-19/USA/WA-S2468/2020\|EPI_ISL_497100\|2020-06-1 | .................... | .................... | .................... | .................... | .................... | .................... |
| F | 2562 hCoV-19/USA/WA-S2469/2020\|EPI_ISL_497101\|2020-06-1 | .................... | .................... | .................... | .................... | .................... | .................... |
| F | 2563 hCoV-19/USA/WA-S2470/2020\|EPI_ISL_497102\|2020-06-1 | .................... | .................... | .................... | .................... | .................... | .................... |
| F | 2564 hCoV-19/USA/WA-S2471/2020\|EPI_ISL_497103\|2020-06-1 | .................... | .................... | .................... | .................... | .................... | .................... |
| F | 2565 hCoV-19/USA/WA-S2472/2020\|EPI_ISL_497104\|2020-06-1 | .................... | .................... | .................... | .................... | .................... | .................... |
| F | 2566 hCoV-19/USA/WA-S2473/2020\|EPI_ISL_497105\|2020-06-1 | .................... | .................... | .................... | .................... | .................... | .................... |
| F | 2567 hCoV-19/USA/WA-S2474/2020\|EPI_ISL_497106\|2020-06-1 | .................... | .................... | .................... | .................... | .................... | .................... |
| F | 2568 hCoV-19/USA/WA-S2475/2020\|EPI_ISL_497107\|2020-06-1 | .................... | .................... | .................... | .................... | .................... | .................... |
| F | 2569 hCoV-19/USA/WA-S2477/2020\|EPI_ISL_497109\|2020-06-1 | .................... | .................... | .................... | .................... | .................... | .................... |
| F | 2570 hCoV-19/USA/WA-S2478/2020\|EPI_ISL_497110\|2020-06-1 | .................... | .................... | .................... | .................... | .................... | .................... |
| F | 2571 hCoV-19/USA/WA-S2479/2020\|EPI_ISL_497111\|2020-06-1 | .................... | .................... | .................... | .................... | .................... | .................... |
| F | 2572 hCoV-19/USA/WA-S2480/2020\|EPI_ISL_497112\|2020-06-1 | .................... | .................... | .................... | .................... | .................... | .................... |
| F | 2573 hCoV-19/USA/WA-S2481/2020\|EPI_ISL_497113\|2020-06-1 | .................... | .................... | .................... | .................... | .................... | .................... |
| F | 2574 hCoV-19/USA/WA-S2482/2020\|EPI_ISL_497114\|2020-06-1 | .................... | .................... | .................... | .................... | .................... | .................... |
| F | 2575 hCoV-19/USA/WA-S2483/2020\|EPI_ISL_497115\|2020-06-1 | .................... | .................... | .................... | .................... | .................... | .................... |
| F | 2576 hCoV-19/USA/WA-S2484/2020\|EPI_ISL_497116\|2020-06-1 | .................... | .................... | .................... | .................... | .................... | .................... |
| F | 2577 hCoV-19/USA/WA-S2485/2020\|EPI_ISL_497117\|2020-06-1 | .................... | .................... | .................... | .................... | .................... | .................... |
| F | 2578 hCoV-19/USA/WA-S2486/2020\|EPI_ISL_497118\|2020-06-1 | .................... | .................... | .................... | .................... | .................... | .................... |
| F | 2579 hCoV-19/USA/WA-S2487/2020\|EPI_ISL_497119\|2020-06-1 | .................... | .................... | .................... | .................... | .................... | .................... |
| F | 2580 hCoV-19/USA/WA-S2488/2020\|EPI_ISL_497120\|2020-06-1 | .................... | .................... | .................... | .................... | .................... | .................... |
| F | 2581 hCoV-19/USA/WA-S2489/2020\|EPI_ISL_497121\|2020-06-1 | .................... | .................... | .................... | .................... | .................... | .................... |
| F | 2582 hCoV-19/USA/WA-S2490/2020\|EPI_ISL_497122\|2020-06-1 | .................... | .................... | .................... | .................... | .................... | .................... |
| F | 2583 hCoV-19/USA/WA-S2491/2020\|EPI_ISL_497123\|2020-06-1 | .................... | .................... | .................... | .................... | .................... | .................... |
| F | 2584 hCoV-19/USA/WA-S2492/2020\|EPI_ISL_497124\|2020-06-1 | .................... | .................... | .................... | .................... | .................... | .................... |
| F | 2585 hCoV-19/USA/WA-S2498/2020\|EPI_ISL_497130\|2020-06-1 | .................... | .................... | .................... | .................... | .................... | .................... |
| F | 2586 hCoV-19/USA/WA-S2499/2020\|EPI_ISL_497131\|2020-06-1 | .................... | .................... | .................... | .................... | .................... | .................... |
| F | 2587 hCoV-19/USA/WA-S2500/2020\|EPI_ISL_497132\|2020-06-1 | .................... | .................... | .................... | .................... | .................... | .................... |
| F | 2588 hCoV-19/USA/WA-S2501/2020\|EPI_ISL_497133\|2020-06-1 | .................... | .................... | .................... | .................... | .................... | .................... |
| F | 2589 hCoV-19/USA/WA-S2502/2020\|EPI_ISL_497134\|2020-06-1 | .................... | .................... | .................... | .................... | .................... | .................... |
| F | 2590 hCoV-19/USA/WA-S2503/2020\|EPI_ISL_497135\|2020-06-1 | .................... | .................... | .................... | .................... | .................... | .................... |
| F | 2591 hCoV-19/USA/WA-S2515/2020\|EPI_ISL_497147\|2020-06-1 | .................... | .................... | .................... | .................... | .................... | .................... |
| F | 2592 hCoV-19/USA/WA-S2516/2020\|EPI_ISL_497148\|2020-06-1 | .................... | .................... | .................... | .................... | .................... | .................... |
| F | 2593 hCoV-19/USA/WA-S2517/2020\|EPI_ISL_497149\|2020-06-1 | .................... | .................... | .................... | .................... | .................... | .................... |
| F | 2594 hCoV-19/USA/WA-S2518/2020\|EPI_ISL_497150\|2020-06-1 | .................... | .................... | .................... | .................... | .................... | .................... |
| F | 2595 hCoV-19/USA/WA-S2519/2020\|EPI_ISL_497151\|2020-06-1 | .................... | .................... | .................... | .................... | .................... | .................... |
| F | 2596 hCoV-19/USA/WA-S2520/2020\|EPI_ISL_497152\|2020-06-1 | .................... | .................... | .................... | .................... | .................... | .................... |
| F | 2597 hCoV-19/USA/WA-S2522/2020\|EPI_ISL_497154\|2020-06-1 | .................... | .................... | .................... | .................... | .................... | .................... |
| F | 2598 hCoV-19/USA/WA-S2523/2020\|EPI_ISL_497155\|2020-06-1 | .................... | .................... | .................... | .................... | .................... | .................... |
| F | 2599 hCoV-19/USA/WA-S2524/2020\|EPI_ISL_497156\|2020-06-1 | .................... | .................... | .................... | .................... | .................... | .................... |
| F | 2600 hCoV-19/USA/WA-S2525/2020\|EPI_ISL_497157\|2020-06-1 | .................... | .................... | .................... | .................... | .................... | .................... |
| F | 2601 hCoV-19/USA/WA-S2526/2020\|EPI_ISL_497158\|2020-06-1 | .................... | .................... | .................... | .................... | .................... | .................... |
| F | 2602 hCoV-19/USA/WA-S2527/2020\|EPI_ISL_497159\|2020-06-1 | .................... | .................... | .................... | .................... | .................... | .................... |
| F | 2603 hCoV-19/USA/WA-S2528/2020\|EPI_ISL_497160\|2020-06-1 | .................... | .................... | .................... | .................... | .................... | .................... |
| F | 2604 hCoV-19/USA/WA-S2529/2020\|EPI_ISL_497161\|2020-06-1 | .................... | .................... | .................... | .................... | .................... | .................... |
| F | 2605 hCoV-19/USA/WA-S2530/2020\|EPI_ISL_497162\|2020-06-1 | .................... | .................... | .................... | .................... | .................... | .................... |
| F | 2606 hCoV-19/USA/WA-S2531/2020\|EPI_ISL_497163\|2020-06-1 | .................... | .................... | .................... | .................... | .................... | .................... |
| F | 2607 hCoV-19/USA/WA-S2532/2020\|EPI_ISL_497164\|2020-06-1 | .................... | .................... | .................... | .................... | .................... | .................... |
| F | 2608 hCoV-19/USA/WA-S2533/2020\|EPI_ISL_497165\|2020-06-1 | .................... | .................... | .................... | .................... | .................... | .................... |
| F | 2609 hCoV-19/USA/WA-S2534/2020\|EPI_ISL_497166\|2020-06-1 | .................... | .................... | .................... | .................... | .................... | .................... |
| F | 2610 hCoV-19/USA/WA-S2536/2020\|EPI_ISL_497168\|2020-06-1 | .................... | .................... | .................... | .................... | .................... | .................... |
| F | 2611 hCoV-19/Bangladesh/BCSIR-NILMRC-276/2020\|EPI_ISL_4 | .................... | .................... | .................... | .................... | .................... | .................... |
| F | 2612 hCoV-19/Bangladesh/BCSIR-NILMRC-280/2020\|EPI_ISL_4 | .................... | .................... | .................... | .................... | .................... | .................... |
| F | 2613 hCoV-19/Bangladesh/BCSIR-NILMRC-282/2020\|EPI_ISL_4 | .................... | .................... | .................... | .................... | .................... | .................... |
| F | 2614 hCoV-19/Australia/ACT0011/2020\|EPI_ISL_498478\|2020 | .................... | .................... | .................... | .................... | .................... | .................... |
| F | 2615 hCoV-19/Australia/ACT0013/2020\|EPI_ISL_498480\|2020 | .................... | .................... | .................... | .................... | .................... | .................... |
| F | 2616 hCoV-19/Australia/ACT0016/2020\|EPI_ISL_498483\|2020 | .................... | .................... | .................... | .................... | .................... | .................... |
| F | 2617 hCoV-19/Australia/ACT0018/2020\|EPI_ISL_498485\|2020 | .................... | .................... | .................... | .................... | .................... | .................... |
| F | 2618 hCoV-19/Australia/ACT0019/2020\|EPI_ISL_498486\|2020 | .................... | .................... | .................... | .................... | .................... | .................... |
| F | 2619 hCoV-19/Australia/ACT0022/2020\|EPI_ISL_498489\|2020 | .................... | .................... | .................... | .................... | .................... | .................... |
| F | 2620 hCoV-19/Australia/ACT0023/2020\|EPI_ISL_498490\|2020 | .................... | .................... | .................... | .................... | .................... | .................... |
| F | 2621 hCoV-19/Australia/ACT0024/2020\|EPI_ISL_498491\|2020 | .................... | .................... | .................... | .................... | .................... | .................... |
| F | 2622 hCoV-19/Australia/ACT0025/2020\|EPI_ISL_498492\|2020 | .................... | .................... | .................... | .................... | .................... | .................... |
| F | 2623 hCoV-19/Australia/ACT0026/2020\|EPI_ISL_498493\|2020 | .................... | .................... | .................... | .................... | .................... | .................... |
| F | 2624 hCoV-19/Australia/ACT0027/2020\|EPI_ISL_498494\|2020 | .................... | .................... | .................... | .................... | .................... | .................... |
| F | 2625 hCoV-19/Australia/ACT0028/2020\|EPI_ISL_498495\|2020 | .................... | .................... | .................... | .................... | .................... | .................... |
| F | 2626 hCoV-19/Australia/ACT0029/2020\|EPI_ISL_498496\|2020 | .................... | .................... | .................... | .................... | .................... | .................... |
| F | 2627 hCoV-19/Australia/ACT0030/2020\|EPI_ISL_498497\|2020 | .................... | .................... | .................... | .................... | .................... | .................... |
| F | 2628 hCoV-19/Australia/ACT0031/2020\|EPI_ISL_498498\|2020 | .................... | .................... | .................... | .................... | .................... | .................... |

| | | | | | | | | | |
|---|---|---|---|---|---|---|---|---|---|
| F | 2629 | hCoV-19/Australia/ACT0032/2020\|EPI_ISL_498499\|2020 | .................... | .................... | .................... | .................... | .................... | .................... | |
| F | 2630 | hCoV-19/England/LIVE-A6303/2020\|EPI_ISL_499700\|202 | .................... | .................... | .................... | .................... | .................... | .................... | |
| F | 2631 | hCoV-19/England/LIVE-A6312/2020\|EPI_ISL_499701\|202 | .................... | .................... | .................... | .................... | .................... | .................... | |
| F | 2632 | hCoV-19/England/LIVE-A635E/2020\|EPI_ISL_499702\|202 | .................... | .................... | .................... | .................... | .................... | .................... | |
| F | 2633 | hCoV-19/England/LIVE-A638B/2020\|EPI_ISL_499703\|202 | .................... | .................... | .................... | .................... | .................... | .................... | |
| F | 2634 | hCoV-19/England/LIVE-A63A9/2020\|EPI_ISL_499704\|202 | .................... | .................... | .................... | .................... | .................... | .................... | |
| F | 2635 | hCoV-19/England/LIVE-A63E5/2020\|EPI_ISL_499705\|202 | .................... | .................... | .................... | .................... | ......NNN........... | .................... | 1 |
| F | 2636 | hCoV-19/England/LIVE-A63F4/2020\|EPI_ISL_499706\|202 | .................... | ......A............. | .................... | .................... | .................... | .................... | |
| F | 2637 | hCoV-19/England/LIVE-A6400/2020\|EPI_ISL_499707\|202 | .................... | .................... | .................... | .................... | .................... | .................... | |
| F | 2638 | hCoV-19/England/LIVE-A643D/2020\|EPI_ISL_499708\|202 | .................... | .................... | .................... | .................... | .................... | .................... | |
| F | 2639 | hCoV-19/England/LIVE-A644C/2020\|EPI_ISL_499709\|202 | .................... | .................... | .................... | .................... | .................... | .................... | |
| F | 2640 | hCoV-19/England/LIVE-A646A/2020\|EPI_ISL_499710\|202 | .................... | .................... | .................... | .................... | .................... | .................... | |
| F | 2641 | hCoV-19/England/LIVE-A6488/2020\|EPI_ISL_499711\|202 | .................... | .................... | .................... | .................... | .................... | .................... | |
| F | 2642 | hCoV-19/England/LIVE-A6497/2020\|EPI_ISL_499712\|202 | .................... | .................... | .................... | .................... | .................... | .................... | |
| F | 2643 | hCoV-19/England/LIVE-A64B5/2020\|EPI_ISL_499713\|202 | .................... | .................... | .................... | .................... | .................... | .................... | |
| F | 2644 | hCoV-19/England/LIVE-A64C4/2020\|EPI_ISL_499714\|202 | .................... | .................... | .................... | .................... | .................... | .................... | |
| F | 2645 | hCoV-19/England/LIVE-A64D3/2020\|EPI_ISL_499715\|202 | .................... | .................... | .................... | .................... | .................... | .................... | |
| F | 2646 | hCoV-19/England/LIVE-A64F1/2020\|EPI_ISL_499716\|202 | .................... | .................... | .................... | .................... | .................... | .................... | |
| F | 2647 | hCoV-19/England/LIVE-A650D/2020\|EPI_ISL_499717\|202 | .................... | .................... | .................... | .................... | .................... | .................... | |
| F | 2648 | hCoV-19/England/LIVE-A653A/2020\|EPI_ISL_499718\|202 | .................... | .................... | .................... | .................... | .................... | .................... | |
| F | 2649 | hCoV-19/England/LIVE-A6549/2020\|EPI_ISL_499719\|202 | .................... | .................... | .................... | .................... | .................... | .................... | |
| F | 2650 | hCoV-19/England/LIVE-A6567/2020\|EPI_ISL_499720\|202 | .................... | .................... | .................... | .................... | .................... | .................... | |
| F | 2652 | hCoV-19/England/LIVE-A6646/2020\|EPI_ISL_499726\|202 | .................... | .................... | .................... | .................... | .................... | .................... | |
| F | 2653 | hCoV-19/England/LIVE-A6655/2020\|EPI_ISL_499727\|202 | .................... | .................... | .................... | .................... | .................... | .................... | |
| F | 2654 | hCoV-19/England/LIVE-A6673/2020\|EPI_ISL_499728\|202 | .................... | .................... | .................... | .................... | .................... | .................... | |
| F | 2655 | hCoV-19/England/LIVE-A66DD/2020\|EPI_ISL_499732\|202 | .................... | .................... | .................... | .................... | .................... | .................... | |
| F | 2656 | hCoV-19/England/LIVE-A66EC/2020\|EPI_ISL_499733\|202 | .................... | .................... | .................... | .................... | .................... | .................... | |
| F | 2657 | hCoV-19/England/LIVE-A66FB/2020\|EPI_ISL_499734\|202 | .................... | .................... | .................... | .................... | .................... | .................... | |
| F | 2658 | hCoV-19/England/LIVE-A6707/2020\|EPI_ISL_499735\|202 | .................... | .................... | .................... | .................... | .................... | .................... | |
| F | 2659 | hCoV-19/England/LIVE-A6734/2020\|EPI_ISL_499736\|202 | .................... | .................... | .................... | .................... | .................... | .................... | |
| F | 2660 | hCoV-19/England/LIVE-A6743/2020\|EPI_ISL_499737\|202 | .................... | .................... | .................... | .................... | .................... | .................... | |
| F | 2661 | hCoV-19/England/LIVE-A6752/2020\|EPI_ISL_499738\|202 | .................... | .................... | .................... | .................... | .................... | .................... | |
| F | 2662 | hCoV-19/England/LIVE-A6761/2020\|EPI_ISL_499739\|202 | .................... | .................... | .................... | .................... | .................... | .................... | |
| F | 2663 | hCoV-19/England/LIVE-A679E/2020\|EPI_ISL_499740\|202 | .................... | .................... | .................... | .................... | .................... | .................... | |
| F | 2664 | hCoV-19/England/LIVE-A67AD/2020\|EPI_ISL_499741\|202 | .................... | .................... | .................... | .................... | .................... | .................... | |
| F | 2665 | hCoV-19/England/LIVE-A67DA/2020\|EPI_ISL_499742\|202 | .................... | .................... | .................... | .................... | .................... | .................... | |
| F | 2666 | hCoV-19/England/LIVE-A67E9/2020\|EPI_ISL_499743\|202 | .................... | .................... | .................... | .................... | .................... | .................... | |
| F | 2667 | hCoV-19/England/LIVE-A67F8/2020\|EPI_ISL_499744\|202 | .................... | .................... | .................... | .................... | .................... | .................... | |
| F | 2668 | hCoV-19/England/LIVE-A6813/2020\|EPI_ISL_499745\|202 | .................... | .................... | .................... | .................... | .................... | .................... | |
| F | 2669 | hCoV-19/England/LIVE-A6831/2020\|EPI_ISL_499746\|202 | .................... | .................... | .................... | .................... | .................... | .................... | |
| F | 2670 | hCoV-19/England/LIVE-A686E/2020\|EPI_ISL_499747\|202 | .................... | .................... | .................... | .................... | .................... | .................... | |
| F | 2671 | hCoV-19/England/LIVE-A687D/2020\|EPI_ISL_499748\|202 | .................... | .................... | .................... | .................... | .................... | .................... | |
| F | 2672 | hCoV-19/England/LIVE-A688C/2020\|EPI_ISL_499749\|202 | .................... | .................... | .................... | .................... | .................... | .................... | |
| F | 2673 | hCoV-19/England/LIVE-A689B/2020\|EPI_ISL_499750\|202 | .................... | .................... | .................... | .................... | .................... | .................... | |
| F | 2674 | hCoV-19/England/LIVE-A68AA/2020\|EPI_ISL_499751\|202 | .................... | .................... | .................... | .................... | .................... | .................... | |
| F | 2675 | hCoV-19/England/LIVE-A68B9/2020\|EPI_ISL_499752\|202 | .................... | .................... | .................... | .................... | .................... | .................... | |
| F | 2676 | hCoV-19/England/LIVE-A68E6/2020\|EPI_ISL_499753\|202 | .................... | .................... | .................... | .................... | .................... | .................... | |
| F | 2677 | hCoV-19/England/LIVE-A68F5/2020\|EPI_ISL_499754\|202 | .................... | ...................A | .................... | .................... | .................... | .................... | 1 |
| F | 2678 | hCoV-19/England/LIVE-A6901/2020\|EPI_ISL_499755\|202 | .................... | .................... | .................... | .................... | .................... | .................... | |
| F | 2680 | hCoV-19/England/LIVE-A695C/2020\|EPI_ISL_499757\|202 | .................... | .................... | .................... | .................... | .................... | .................... | |
| F | 2681 | hCoV-19/England/LIVE-A696B/2020\|EPI_ISL_499758\|202 | .................... | .................... | .................... | .................... | .................... | .................... | |
| F | 2682 | hCoV-19/England/LIVE-A6998/2020\|EPI_ISL_499759\|202 | .................... | .................... | .................... | .................... | .................... | .................... | |
| F | 2683 | hCoV-19/England/LIVE-A69C5/2020\|EPI_ISL_499760\|202 | .................... | .................... | .................... | .................... | .................... | .................... | |
| F | 2684 | hCoV-19/England/LIVE-A69D4/2020\|EPI_ISL_499761\|202 | .................... | .................... | .................... | .................... | .................... | .................... | |
| F | 2685 | hCoV-19/England/LIVE-A69E3/2020\|EPI_ISL_499762\|202 | .................... | .................... | .................... | .................... | .................... | .................... | |
| F | 2686 | hCoV-19/England/LIVE-A69F2/2020\|EPI_ISL_499763\|202 | .................... | .................... | .................... | .................... | .................... | .................... | |
| F | 2687 | hCoV-19/England/LIVE-A6A0E/2020\|EPI_ISL_499764\|202 | .................... | ............A....... | .................... | .................... | .................... | .................... | |
| F | 2688 | hCoV-19/England/LIVE-A6A2C/2020\|EPI_ISL_499765\|202 | .................... | .................... | .................... | .................... | .................... | .................... | |
| F | 2689 | hCoV-19/England/LIVE-A6A4A/2020\|EPI_ISL_499766\|202 | .................... | .................... | .................... | .................... | .................... | .................... | |
| F | 2690 | hCoV-19/England/LIVE-A6A59/2020\|EPI_ISL_499767\|202 | .................... | .................... | .................... | .................... | .................... | .................... | |
| F | 2691 | hCoV-19/England/LIVE-A6A77/2020\|EPI_ISL_499768\|202 | .................... | .................... | .................... | .................... | .................... | .................... | |
| F | 2692 | hCoV-19/England/LIVE-A6AB3/2020\|EPI_ISL_499769\|202 | .................... | ...................A | .................... | .................... | .................... | .................... | 1 |
| F | 2693 | hCoV-19/England/LIVE-A6AD1/2020\|EPI_ISL_499770\|202 | .................... | .................... | .................... | .................... | .................... | .................... | |
| F | 2694 | hCoV-19/England/LIVE-A6AE0/2020\|EPI_ISL_499771\|202 | .................... | .................... | .................... | .................... | .................... | .................... | |
| F | 2695 | hCoV-19/England/LIVE-A6AFF/2020\|EPI_ISL_499772\|202 | .................... | .................... | .................... | .................... | .................... | .................... | |
| F | 2696 | hCoV-19/England/LIVE-A6B0B/2020\|EPI_ISL_499773\|202 | .................... | .................... | .................... | .................... | .................... | .................... | |
| F | 2698 | hCoV-19/England/LIVE-A6B38/2020\|EPI_ISL_499775\|202 | .................... | .................... | .................... | .................... | .................... | .................... | |
| F | 2699 | hCoV-19/England/LIVE-A6B65/2020\|EPI_ISL_499776\|202 | .................... | .................... | .................... | .................... | .................... | .................... | |
| F | 2700 | hCoV-19/England/NORT-29338F/2020\|EPI_ISL_499777\|20 | .................... | .................... | .................... | .................... | .................... | .................... | |
| F | 2701 | hCoV-19/England/NORT-2934D7/2020\|EPI_ISL_499778\|20 | .................... | .................... | .................... | .................... | .................... | .................... | |
| F | 2702 | hCoV-19/England/NORT-29352F/2020\|EPI_ISL_499779\|20 | .................... | .................... | .................... | .................... | .................... | .................... | |
| F | 2703 | hCoV-19/England/NORT-29361D/2020\|EPI_ISL_499780\|20 | .................... | .................... | .................... | .................... | .................... | .................... | |
| F | 2704 | hCoV-19/England/NORT-293668/2020\|EPI_ISL_499781\|20 | .................... | .................... | .................... | .................... | .................... | .................... | |
| F | 2705 | hCoV-19/England/NORT-2937B0/2020\|EPI_ISL_499782\|20 | .................... | .................... | .................... | .................... | .................... | .................... | |
| F | 2706 | hCoV-19/England/NORT-293853/2020\|EPI_ISL_499783\|20 | .................... | .................... | .................... | .................... | .................... | .................... | |
| F | 2707 | hCoV-19/England/NORT-296F5B/2020\|EPI_ISL_499784\|20 | .................... | .A.................. | .................... | .................... | .................... | .................... | |
| F | 2708 | hCoV-19/England/NORT-297127/2020\|EPI_ISL_499785\|20 | .................... | .................... | .................... | .................... | .................... | .................... | |
| F | 2709 | hCoV-19/England/NORT-29C2F2/2020\|EPI_ISL_499786\|20 | .................... | .................... | .................... | .................... | .................... | .................... | |
| F | 2710 | hCoV-19/England/NORT-29CD1F/2020\|EPI_ISL_499790\|20 | .................... | .................... | .................... | .................... | .................... | .................... | |
| F | 2711 | hCoV-19/England/NORT-29CD2E/2020\|EPI_ISL_499791\|20 | .................... | .................... | .................... | .................... | .................... | .................... | |

| | | | | | | | | |
|---|---|---|---|---|---|---|---|---|
| F | 2712 | hCoV-19/England/NORT-29CDA6/2020\|EPI_ISL_499792\|20 | ···················· | ···················· | ···················· | ···················· | ···················· | ···················· |
| F | 2713 | hCoV-19/England/NORT-29CDB5/2020\|EPI_ISL_499793\|20 | ···················· | ···················· | ···················· | ···················· | ···················· | ···················· |
| F | 2714 | hCoV-19/England/NORT-29CE0D/2020\|EPI_ISL_499794\|20 | ···················· | ···················· | ···················· | ···················· | ···················· | ···················· |
| F | 2715 | hCoV-19/England/NORT-29CE49/2020\|EPI_ISL_499795\|20 | ···················· | ···················· | ···················· | ···················· | ···················· | ···················· |
| F | 2716 | hCoV-19/England/NORT-29CEB2/2020\|EPI_ISL_499796\|20 | ···················· | ···················· | ···················· | ···················· | ···················· | ···················· |
| F | 2717 | hCoV-19/England/NORT-29D042/2020\|EPI_ISL_499797\|20 | ···················· | ···················· | ···················· | ···················· | ···················· | ···················· |
| F | 2718 | hCoV-19/USA/NM-UNM-00331/2020\|EPI_ISL_508100\|2020- | ···················· | ···················· | ···················· | ···················· | ···················· | ···················· |
| F | 2719 | hCoV-19/USA/NM-UNM-00328/2020\|EPI_ISL_508101\|2020- | ···················· | ···················· | ···················· | ···················· | ···················· | ···················· |
| F | 2720 | hCoV-19/Australia/SAP184/2020\|EPI_ISL_508122\|2020- | ···················· | ···················· | ···················· | ···················· | ···················· | ···················· |
| F | 2721 | hCoV-19/Australia/SAP432/2020\|EPI_ISL_508123\|2020- | ···················· | ···················· | ···················· | ···················· | ···················· | ···················· |
| F | 2722 | hCoV-19/Australia/SAP433/2020\|EPI_ISL_508124\|2020- | ···················· | ···················· | ···················· | ···················· | ···················· | ···················· |
| F | 2723 | hCoV-19/Australia/SAP434/2020\|EPI_ISL_508125\|2020- | ···················· | ·············A······ | ···················· | ···················· | ···················· | ···················· |
| F | 2724 | hCoV-19/Australia/SAP435/2020\|EPI_ISL_508126\|2020- | ···················· | ···················· | ···················· | ···················· | ···················· | ···················· |
| F | 2725 | hCoV-19/Australia/SAP436/2020\|EPI_ISL_508127\|2020- | ···················· | ···················· | ···················· | ···················· | ···················· | ···················· |
| F | 2726 | hCoV-19/India/UT-AR22/2020\|EPI_ISL_508170\|2020-05- | ···················· | ···················· | ···················· | ···················· | ···················· | ···················· |
| F | 2727 | hCoV-19/India/UT-AR23/2020\|EPI_ISL_508171\|2020-05- | ···················· | ···················· | ···················· | ···················· | ···················· | ···················· |
| F | 2728 | hCoV-19/India/UT-AR24/2020\|EPI_ISL_508172\|2020-05- | ···················· | ·A·················· | ···················· | ···················· | ···················· | ··A················· |
| F | 2729 | hCoV-19/India/UT-AR25/2020\|EPI_ISL_508173\|2020-05- | ···················· | ···················· | ···················· | ···················· | ···················· | ···················· |
| F | 2730 | hCoV-19/India/UT-AR27/2020\|EPI_ISL_508174\|2020-06- | ···················· | ···················· | ···················· | ···················· | ···················· | ···················· |
| F | 2731 | hCoV-19/India/UT-AR28/2020\|EPI_ISL_508175\|2020-06- | ···················· | ·A·················· | ···················· | ···················· | ···················· | ··A················· |
| F | 2732 | hCoV-19/India/UT-AR29/2020\|EPI_ISL_508176\|2020-05- | ···················· | ·A·················· | ···················· | ···················· | ···················· | ··A················· |
| F | 2733 | hCoV-19/India/UT-AR3/2020\|EPI_ISL_508177\|2020-06-0 | ···················· | ·A·················· | ···················· | ···················· | ···················· | ··A················· |
| F | 2734 | hCoV-19/India/UT-AR30/2020\|EPI_ISL_508178\|2020-05- | ···················· | ···················· | ···················· | ···················· | ···················· | ···················· |
| F | 2735 | hCoV-19/India/UT-AR31/2020\|EPI_ISL_508179\|2020-05- | ···················· | ···················· | ···················· | ···················· | ···················· | ···················· |
| F | 2736 | hCoV-19/India/UT-AR46/2020\|EPI_ISL_508190\|2020-05- | ···················· | ···················· | ···················· | ···················· | ···················· | ···················· |
| F | 2737 | hCoV-19/India/UT-AR5/2020\|EPI_ISL_508191\|2020-06-0 | ···················· | ···················· | ···················· | ···················· | ···················· | ···················· |
| F | 2738 | hCoV-19/India/UT-AR54/2020\|EPI_ISL_508193\|2020-06- | ···················· | ···················· | ···················· | ···················· | ···················· | ···················· |
| F | 2739 | hCoV-19/India/UT-AR58/2020\|EPI_ISL_508194\|2020-05- | ···················· | ···················· | ···················· | ···················· | ···················· | ···················· |
| F | 2740 | hCoV-19/India/UT-AR59/2020\|EPI_ISL_508195\|2020-06- | ···················· | ·A·················· | ···················· | ···················· | ···················· | ··A················· |
| F | 2741 | hCoV-19/India/UT-AR6/2020\|EPI_ISL_508196\|2020-05-2 | ···················· | ·A·················· | ···················· | ···················· | ···················· | ··A················· |
| F | 2742 | hCoV-19/India/UT-AR60/2020\|EPI_ISL_508197\|2020-06- | ···················· | ···················· | ···················· | ···················· | ···················· | ···················· |
| F | 2743 | hCoV-19/India/UT-AR61/2020\|EPI_ISL_508198\|2020-06- | ···················· | ···················· | ···················· | ···················· | ···················· | ···················· |
| F | 2744 | hCoV-19/India/UT-AR62/2020\|EPI_ISL_508199\|2020-06- | ···················· | ···················· | ···················· | ···················· | ···················· | ···················· |
| F | 2745 | hCoV-19/USA/AL-P44e/2020\|EPI_ISL_509424\|2020-05-12 | ···················· | ···················· | ···················· | ···················· | ···················· | ···················· |
| F | 2746 | hCoV-19/USA/AL-SV100V1/2020\|EPI_ISL_509425\|2020-05 | ···················· | ···················· | ···················· | ···················· | ···················· | ···················· |
| F | 2747 | hCoV-19/USA/AL-SV101V1/2020\|EPI_ISL_509426\|2020-04 | ···················· | ···················· | ···················· | ···················· | ···················· | ···················· |
| F | 2748 | hCoV-19/USA/AL-SV106V1/2020\|EPI_ISL_509427\|2020-05 | ···················· | ···················· | ···················· | ···················· | ···················· | ···················· |
| F | 2749 | hCoV-19/USA/AL-SV107V1/2020\|EPI_ISL_509428\|2020-05 | ···················· | ···················· | ···················· | ···················· | ···················· | ···················· |
| F | 2750 | hCoV-19/USA/AL-SV110BV1/2020\|EPI_ISL_509429\|2020-0 | ···················· | ···················· | ···················· | ···················· | ···················· | ···················· |
| F | 2751 | hCoV-19/Brazil/RJ-DCV05/2020\|EPI_ISL_509430\|2020-0 | ···················· | ···················· | ···················· | ···················· | ···················· | ···················· |
| F | 2752 | hCoV-19/Brazil/RJ-DCV1/2020\|EPI_ISL_509431\|2020-07 | ···················· | ···················· | ···················· | ···················· | ···················· | ···················· |
| F | 2753 | hCoV-19/Brazil/RJ-DCV3/2020\|EPI_ISL_509432\|2020-07 | ···················· | ···················· | ···················· | ···················· | ···················· | ···················· |
| F | 2754 | hCoV-19/Brazil/RJ-DCV5/2020\|EPI_ISL_509433\|2020-07 | ···················· | ···················· | ···················· | ···················· | ···················· | ···················· |
| F | 2755 | hCoV-19/Brazil/RJ-DCVN1/2020\|EPI_ISL_509434\|2020-0 | ···················· | ···················· | ···················· | ···················· | ···················· | ···················· |
| F | 2756 | hCoV-19/Brazil/RJ-DCVTV507Q/2020\|EPI_ISL_509435\|20 | ···················· | ···················· | ···················· | ···················· | ···················· | ···················· |
| F | 2757 | hCoV-19/England/PRIN-254BC91/2020\|EPI_ISL_509436\|2 | ···················· | ···················· | ···················· | ···················· | ···················· | ···················· |
| F | 2758 | hCoV-19/England/PRIN-254BCBF/2020\|EPI_ISL_509439\|2 | ···················· | ···················· | ···················· | ···················· | ···················· | ···················· |
| F | 2759 | hCoV-19/USA/MD-MDH-0046/2020\|EPI_ISL_509450\|2020-0 | ···················· | ···················· | ···················· | ···················· | ···················· | ···················· |
| F | 2760 | hCoV-19/USA/MD-MDH-0047/2020\|EPI_ISL_509451\|2020-0 | ···················· | ···················· | ···················· | ···················· | ···················· | ···················· |
| F | 2761 | hCoV-19/USA/MD-MDH-0048/2020\|EPI_ISL_509452\|2020-0 | ···················· | ···················· | ···················· | ···················· | ···················· | ···················· |
| F | 2762 | hCoV-19/USA/MD-MDH-0049/2020\|EPI_ISL_509453\|2020-0 | ···················· | ···················· | ···················· | ···················· | ···················· | ···················· |
| F | 2763 | hCoV-19/USA/MD-MDH-0050/2020\|EPI_ISL_509454\|2020-0 | ···················· | ···················· | ···················· | ···················· | ···················· | ···················· |
| F | 2764 | hCoV-19/USA/MD-MDH-0051/2020\|EPI_ISL_509455\|2020-0 | ···················· | ···················· | ···················· | ···················· | ···················· | ···················· |
| F | 2765 | hCoV-19/USA/MD-MDH-0052/2020\|EPI_ISL_509456\|2020-0 | ···················· | ···················· | ···················· | ···················· | ···················· | ···················· |
| F | 2766 | hCoV-19/USA/MD-MDH-0053/2020\|EPI_ISL_509457\|2020-0 | ···················· | ···················· | ···················· | ···················· | ···················· | ···················· |
| F | 2767 | hCoV-19/USA/MD-MDH-0054/2020\|EPI_ISL_509458\|2020-0 | ···················· | ···················· | ···················· | ···················· | ···················· | ···················· |
| F | 2770 | hCoV-19/Spain/PV-IBV-004288/2020\|EPI_ISL_510404\|20 | ···················· | ···················· | ···················· | ···················· | ···················· | ···················· |
| F | 2771 | hCoV-19/Spain/PV-IBV-004289/2020\|EPI_ISL_510405\|20 | ···················· | ···················· | ···················· | ···················· | ··NN················ | ···················· |
| F | 2772 | hCoV-19/Spain/PV-IBV-004290/2020\|EPI_ISL_510406\|20 | ···················· | ···················· | ···················· | ···················· | ···················· | ···················· |
| F | 2773 | hCoV-19/Spain/PV-IBV-004291/2020\|EPI_ISL_510407\|20 | ···················· | ···················· | ···················· | ···················· | ···················· | ···················· |
| F | 2789 | hCoV-19/Portugal/PT1188/2020\|EPI_ISL_511733\|2020-0 | ···················· | ···················· | ···················· | ···················· | ···················· | ···················· |
| F | 2790 | hCoV-19/Portugal/PT1197/2020\|EPI_ISL_511741\|2020-0 | ···················· | ···················· | ···················· | ···················· | ···················· | ···················· |
| F | 2791 | hCoV-19/Portugal/PT1205/2020\|EPI_ISL_511748\|2020-0 | ···················· | ···················· | ···················· | ···················· | ···················· | ···················· |
| F | 2792 | hCoV-19/Portugal/PT1209/2020\|EPI_ISL_511752\|2020-0 | ···················· | ···················· | ···················· | ···················· | ···················· | ···················· |
| F | 2793 | hCoV-19/Portugal/PT1210/2020\|EPI_ISL_511753\|2020-0 | ···················· | ···················· | ···················· | ···················· | ···················· | ···················· |
| F | 2794 | hCoV-19/Portugal/PT1211/2020\|EPI_ISL_511754\|2020-0 | ···················· | ···················· | ···················· | ···················· | ···················· | ···················· |
| F | 2795 | hCoV-19/Portugal/PT1217/2020\|EPI_ISL_511760\|2020-0 | ···················· | ···················· | ···················· | ···················· | ···················· | ···················· |
| F | 2796 | hCoV-19/Portugal/PT1218/2020\|EPI_ISL_511761\|2020-0 | ···················· | ···················· | ···················· | ···················· | ···················· | ···················· |
| F | 2797 | hCoV-19/Portugal/PT1219/2020\|EPI_ISL_511762\|2020-0 | ···················· | ···················· | ···················· | ···················· | ···················· | ···················· |
| F | 2798 | hCoV-19/Portugal/PT1220/2020\|EPI_ISL_511763\|2020-0 | ···················· | ···················· | ···················· | ···················· | ···················· | ···················· |
| F | 2799 | hCoV-19/Portugal/PT1221/2020\|EPI_ISL_511764\|2020-0 | ···················· | ···················· | ···················· | ···················· | ···················· | ···················· |
| F | 2800 | hCoV-19/Portugal/PT1222/2020\|EPI_ISL_511765\|2020-0 | ···················· | ···················· | ···················· | ···················· | ···················· | ···················· |
| F | 2801 | hCoV-19/Switzerland/GE-ETHZ-230065/2020\|EPI_ISL_51 | ···················· | ···················· | ···················· | ···················· | ···················· | ···················· |
| F | 2802 | hCoV-19/Switzerland/VD-ETHZ-230066/2020\|EPI_ISL_51 | ···················· | ···················· | ···················· | ···················· | ···················· | ···················· |
| F | 2803 | hCoV-19/Switzerland/ZH-ETHZ-230069/2020\|EPI_ISL_51 | ···················· | ···················· | ···················· | ···················· | ···················· | ···················· |
| F | 2804 | hCoV-19/Switzerland/ZH-ETHZ-230070/2020\|EPI_ISL_51 | ···················· | ···················· | ···················· | ···················· | ···················· | ···················· |
| F | 2805 | hCoV-19/Switzerland/ZH-ETHZ-230073/2020\|EPI_ISL_51 | ···················· | ···················· | ···················· | ···················· | ···················· | ···················· |
| F | 2806 | hCoV-19/Mexico/BCN-InDRE-45/2020\|EPI_ISL_516609\|20 | ···················· | ·A·················· | ···················· | ···················· | ···················· | ···················· |
| F | 2807 | hCoV-19/Mexico/BCN-InDRE-51/2020\|EPI_ISL_516611\|20 | ···················· | ·A·················· | ···················· | ···················· | ···················· | ···················· |
| F | 2808 | hCoV-19/Mexico/CHH-InDRE-43/2020\|EPI_ISL_516613\|20 | ···················· | ···················· | ···················· | ···················· | ···················· | ···················· |

| | | | | | | | |
|---|---|---|---|---|---|---|---|
| F | 2809 hCoV-19/Mexico/COA-InDRE-36/2020\|EPI_ISL_516618\|20 | ···················· | ···················· | ···················· | ···················· | ···················· | ···················· |
| F | 2810 hCoV-19/Mexico/COA-InDRE_40/2020\|EPI_ISL_516620\|20 | ···················· | ···················· | ···················· | ···················· | ···················· | ···················· |
| F | 2811 hCoV-19/Mexico/SON-InDRE_35/2020\|EPI_ISL_516622\|20 | ···················· | ·A·················· | ···················· | ···················· | ···················· | ···················· |
| F | 2812 hCoV-19/Mexico/TAM-InDRE_38/2020\|EPI_ISL_516624\|20 | ···················· | ···················· | ···················· | ···················· | ···················· | ···················· |
| F | 2813 hCoV-19/Mexico/TAM-InDRE_39/2020\|EPI_ISL_516625\|20 | ···················· | ···················· | ···················· | ···················· | ···················· | ···················· |
| F | 2814 hCoV-19/Peru/ARE-INS-087/2020\|EPI_ISL_516626\|2020- | ···················· | ···················· | ···················· | ···················· | ···················· | ···················· |
| F | 2815 hCoV-19/Estonia/ChVir4307/2020\|EPI_ISL_516627\|2020 | ···················· | ···················· | ···················· | ···················· | ···················· | ···················· |
| F | 2816 hCoV-19/Germany/BY-UHR-20/2020\|EPI_ISL_522407\|2020 | ···················· | ···················· | ···················· | ···················· | ···················· | ···················· |
| F | 2817 hCoV-19/England/LIVE-A6576/2020\|EPI_ISL_499721\|202 | ···················· | ···················· | ···················· | ···················· | ···················· | ···················· |
| F | 2818 hCoV-19/England/LIVE-A6594/2020\|EPI_ISL_499722\|202 | ···················· | ···················· | ···················· | ···················· | ···················· | ···················· |
| F | 2819 hCoV-19/England/LIVE-A65B2/2020\|EPI_ISL_499723\|202 | ···················· | ···················· | ···················· | ···················· | ···················· | ···················· |
| F | 2820 hCoV-19/England/LIVE-A65EF/2020\|EPI_ISL_499724\|202 | ···················· | ···················· | ···················· | ···················· | ···················· | ···················· |
| F | 2821 hCoV-19/England/LIVE-A6691/2020\|EPI_ISL_499729\|202 | ···················· | ···················· | ···················· | ···················· | ···················· | ···················· |
| F | 2822 hCoV-19/England/LIVE-A66A0/2020\|EPI_ISL_499730\|202 | ···················· | ···················· | ···················· | ···················· | ···················· | ···················· |
| F | 2823 hCoV-19/England/LIVE-A66CE/2020\|EPI_ISL_499731\|202 | ···················· | ···················· | ···················· | ···················· | ···················· | ···················· |
| F | 2824 hCoV-19/India/UT-AR32/2020\|EPI_ISL_508180\|2020-06- | ···················· | ·A·················· | ···················· | ···················· | ···················· | ··A················· |
| F | 2825 hCoV-19/India/UT-AR33/2020\|EPI_ISL_508181\|2020-06- | ···················· | ···················· | ···················· | ···················· | ···················· | ···················· |
| F | 2826 hCoV-19/India/UT-AR34/2020\|EPI_ISL_508182\|2020-06- | ···················· | ·A·················· | ···················· | ···················· | ···················· | ··A················· |
| F | 2827 hCoV-19/India/UT-AR36/2020\|EPI_ISL_508183\|2020-05- | ···················· | ···················· | ···················· | ···················· | ···················· | ···················· |
| F | 2828 hCoV-19/India/UT-AR4/2020\|EPI_ISL_508184\|2020-06-0 | ···················· | ···················· | ···················· | ···················· | ···················· | ···················· |
| F | 2829 hCoV-19/India/UT-AR41/2020\|EPI_ISL_508185\|2020-05- | ···················· | ···················· | ···················· | ···················· | ···················· | ···················· |
| F | 2830 hCoV-19/India/UT-AR42/2020\|EPI_ISL_508186\|2020-05- | ···················· | ···················· | ···················· | ···················· | ···················· | ···················· |
| F | 2831 hCoV-19/India/UT-AR43/2020\|EPI_ISL_508187\|2020-05- | ···················· | ···················· | ···················· | ···················· | ···················· | ···················· |
| F | 2832 hCoV-19/India/UT-AR44/2020\|EPI_ISL_508188\|2020-05- | ···················· | ···················· | ···················· | ···················· | ···················· | ···················· |
| F | 2833 hCoV-19/India/UT-AR45/2020\|EPI_ISL_508189\|2020-05- | ···················· | ···················· | ···················· | ···················· | ···················· | ···················· |
| F | 2834 hCoV-19/India/UT-AR63/2020\|EPI_ISL_508200\|2020-06- | ···················· | ·A·················· | ···················· | ···················· | ···················· | ··A················· |
| F | 2835 hCoV-19/India/UT-AR65/2020\|EPI_ISL_508201\|2020-06- | ···················· | ···················· | ···················· | ···················· | ···················· | ···················· |
| F | 2836 hCoV-19/India/UP-AR66/2020\|EPI_ISL_508202\|2020-06- | ···················· | ·A·················· | ···················· | ···················· | ···················· | ··A················· |
| F | 2837 hCoV-19/India/UP-AR67/2020\|EPI_ISL_508203\|2020-06- | ···················· | ·A·················· | ···················· | ···················· | ···················· | ··A················· |
| F | 2838 hCoV-19/India/UT-AR7/2020\|EPI_ISL_508204\|2020-05-3 | ···················· | ···················· | ···················· | ···················· | ···················· | ···················· |
| F | 2839 hCoV-19/India/UT-AR8/2020\|EPI_ISL_508205\|2020-05-2 | ···················· | ···················· | ···················· | ···················· | ···················· | ···················· |
| F | 2840 hCoV-19/India/UT-AR9/2020\|EPI_ISL_508206\|2020-05-2 | ···················· | ···················· | ···················· | ···················· | ···················· | ···················· |
| F | 2841 hCoV-19/India/MH-GA11/2020\|EPI_ISL_508207\|2020-06- | ···················· | ···················· | ···················· | ···················· | ···················· | ···················· |
| F | 2842 hCoV-19/India/MH-GA12/2020\|EPI_ISL_508208\|2020-06- | ···················· | ···················· | ···················· | ···················· | ···················· | ···················· |
| F | 2843 hCoV-19/India/MH-GA13/2020\|EPI_ISL_508209\|2020-06- | ···················· | ···················· | ···················· | ···················· | ···················· | ···················· |
| F | 2844 hCoV-19/Australia/NSW2126/2020\|EPI_ISL_509500\|2020 | ···················· | ···················· | ···················· | ···················· | ···················· | ···················· |
| F | 2845 hCoV-19/Australia/NSW2142/2020\|EPI_ISL_509501\|2020 | ···················· | ···················· | ···················· | ···················· | ···················· | ···················· |
| F | 2846 hCoV-19/Australia/NSW2143/2020\|EPI_ISL_509502\|2020 | ···················· | ···················· | ···················· | ···················· | ···················· | ···················· |
| F | 2847 hCoV-19/Australia/NSW2146/2020\|EPI_ISL_509503\|2020 | ···················· | ···················· | ···················· | ···················· | ···················· | ···················· |
| F | 2848 hCoV-19/Australia/NSW2152/2020\|EPI_ISL_509504\|2020 | ···················· | ···················· | ···················· | ···················· | ···················· | ···················· |
| F | 2849 hCoV-19/Australia/NSW2153/2020\|EPI_ISL_509505\|2020 | ···················· | ···················· | ···················· | ···················· | ···················· | ···················· |
| F | 2850 hCoV-19/Australia/NSW2166/2020\|EPI_ISL_509506\|2020 | ···················· | ···················· | ···················· | ···················· | ···················· | ···················· |
| F | 2851 hCoV-19/Australia/NSW2177/2020\|EPI_ISL_509507\|2020 | ···················· | ···················· | ···················· | ···················· | ···················· | ···················· |
| F | 2852 hCoV-19/Australia/NSW2180/2020\|EPI_ISL_509508\|2020 | ···················· | ···················· | ···················· | ···················· | ···················· | ···················· |
| F | 2853 hCoV-19/Australia/NSW2182/2020\|EPI_ISL_509509\|2020 | ···················· | ···················· | ···················· | ···················· | ···················· | ···················· |
| F | 2854 hCoV-19/Australia/NSW2235/2020\|EPI_ISL_509514\|2020 | ···················· | ···················· | ···················· | ···················· | ···················· | ···················· |
| F | 2855 hCoV-19/Australia/NSW2236/2020\|EPI_ISL_509515\|2020 | ···················· | ···················· | ···················· | ···················· | ···················· | ···················· |
| F | 2856 hCoV-19/Australia/NSW2241/2020\|EPI_ISL_509516\|2020 | ···················· | ···················· | ···················· | ···················· | ···················· | ···················· |
| F | 2857 hCoV-19/Australia/NSW2273/2020\|EPI_ISL_509517\|2020 | ···················· | ···················· | ···················· | ···················· | ···················· | ···················· |
| F | 2858 hCoV-19/Australia/NSW2318/2020\|EPI_ISL_509520\|2020 | ···················· | ···················· | ···················· | ···················· | ···················· | ···················· |
| F | 2859 hCoV-19/Australia/NSW2520/2020\|EPI_ISL_509521\|2020 | ···················· | ···················· | ···················· | ···················· | ···················· | ···················· |
| F | 2860 hCoV-19/Australia/NSW2530/2020\|EPI_ISL_509522\|2020 | ···················· | ···················· | ···················· | ···················· | ···················· | ···················· |
| F | 2861 hCoV-19/Australia/NSW2579/2020\|EPI_ISL_509523\|2020 | ···················· | ···················· | ···················· | ···················· | ···················· | ···················· |
| F | 2862 hCoV-19/USA/UT-02883/2020\|EPI_ISL_509528\|2020-04-3 | ···················· | ···················· | ···················· | ···················· | ···················· | ···················· |
| F | 2863 hCoV-19/USA/UT-02884/2020\|EPI_ISL_509529\|2020-04-3 | ···················· | ···················· | ···················· | ···················· | ···················· | ···················· |
| F | 2864 hCoV-19/USA/UT-02885/2020\|EPI_ISL_509530\|2020-04-3 | ···················· | ···················· | ···················· | ···················· | ···················· | ···················· |
| F | 2865 hCoV-19/USA/UT-02886/2020\|EPI_ISL_509531\|2020-04-3 | ···················· | ···················· | ···················· | ···················· | ···················· | ···················· |
| F | 2866 hCoV-19/USA/UT-02903/2020\|EPI_ISL_509534\|2020-04-3 | ···················· | ···················· | ···················· | ···················· | ···················· | ···················· |
| F | 2867 hCoV-19/USA/UT-02904/2020\|EPI_ISL_509535\|2020-04-3 | ···················· | ···················· | ···················· | ···················· | ···················· | ···················· |
| F | 2868 hCoV-19/USA/UT-02909/2020\|EPI_ISL_509536\|2020-04-3 | ···················· | ···················· | ···················· | ···················· | ···················· | ···················· |
| F | 2869 hCoV-19/USA/UT-02910/2020\|EPI_ISL_509537\|2020-04-3 | ···················· | ···················· | ···················· | ···················· | ···················· | ···················· |
| F | 2879 hCoV-19/Spain/PV-IBV-004215/2020\|EPI_ISL_510500\|20 | ···················· | ···················· | ···················· | ···················· | ···················· | ···················· |
| F | 2882 hCoV-19/Spain/PV-IBV-004219/2020\|EPI_ISL_510503\|20 | ···················· | ···················· | ···················· | ···················· | ···················· | ···················· |
| F | 2886 hCoV-19/Spain/PV-IBV-004225/2020\|EPI_ISL_510507\|20 | ···················· | ···················· | ···················· | ···················· | ···················· | ···················· |
| F | 2888 hCoV-19/Spain/PV-IBV-004227/2020\|EPI_ISL_510509\|20 | ···················· | ···················· | ···················· | ···················· | ···················· | ···················· |
| F | 2896 hCoV-19/Spain/CT-IBV-005489/2020\|EPI_ISL_510517\|20 | ···················· | ···················· | ···················· | ···················· | ···················· | ···················· |
| F | 2898 hCoV-19/Spain/CT-IBV-005502/2020\|EPI_ISL_510520\|20 | ···················· | ·A·················· | ···················· | ···················· | ···················· | ···················· |
| F | 2900 hCoV-19/USA/CA-IGI-0055/2020\|EPI_ISL_511810\|2020-0 | ···················· | ···················· | ···················· | ···················· | ···················· | ···················· |
| F | 2901 hCoV-19/USA/CA-IGI-0057/2020\|EPI_ISL_511812\|2020-0 | ···················· | ···················· | ···················· | ···················· | ···················· | ···················· |
| F | 2902 hCoV-19/USA/CA-IGI-0059/2020\|EPI_ISL_511814\|2020-0 | ···················· | ···················· | ···················· | ···················· | ···················· | ···················· |
| F | 2903 hCoV-19/USA/CA-IGI-0060/2020\|EPI_ISL_511815\|2020-0 | ···················· | ···················· | ···················· | ···················· | ···················· | ···················· |
| F | 2904 hCoV-19/USA/CA-IGI-0061/2020\|EPI_ISL_511816\|2020-0 | ···················· | ···················· | ···················· | ···················· | ···················· | ···················· |
| F | 2905 hCoV-19/USA/CA-IGI-0062/2020\|EPI_ISL_511817\|2020-0 | ···················· | ···················· | ···················· | ···················· | ···················· | ···················· |
| F | 2906 hCoV-19/USA/CA-IGI-0067/2020\|EPI_ISL_511821\|2020-0 | ···················· | ···················· | ···················· | ···················· | ···················· | ···················· |
| F | 2907 hCoV-19/USA/CA-IGI-0068/2020\|EPI_ISL_511822\|2020-0 | ···················· | ···················· | ···················· | ···················· | ···················· | ···················· |
| F | 2908 hCoV-19/USA/CA-IGI-0069/2020\|EPI_ISL_511823\|2020-0 | ···················· | ·A·················· | ···················· | ···················· | ···················· | ···················· |
| F | 2909 hCoV-19/USA/CA-IGI-0070/2020\|EPI_ISL_511824\|2020-0 | ···················· | ···················· | ···················· | ···················· | ···················· | ···················· |
| F | 2910 hCoV-19/USA/CA-IGI-0071/2020\|EPI_ISL_511825\|2020-0 | ···················· | ·A·················· | ···················· | ···················· | ···················· | ···················· |
| F | 2911 hCoV-19/USA/CA-IGI-0074/2020\|EPI_ISL_511828\|2020-0 | ···················· | ···················· | ···················· | ···················· | ···················· | ···················· |
| F | 2912 hCoV-19/USA/CA-IGI-0075/2020\|EPI_ISL_511829\|2020-0 | ···················· | ···················· | ···················· | ···················· | ···················· | ···················· |

| | | | | | | | |
|---|---|---|---|---|---|---|---|
| F | 2913 hCoV-19/USA/CA-IGI-0076/2020\|EPI_ISL_511830\|2020-0 | .................... | .................... | ..................... | .................... | .................... | .................... |
| F | 2914 hCoV-19/USA/CA-IGI-0077/2020\|EPI_ISL_511831\|2020-0 | .................... | .................... | ..................... | .................... | .................... | .................... |
| F | 2915 hCoV-19/USA/CA-IGI-0078/2020\|EPI_ISL_511832\|2020-0 | .................... | .................... | ..................... | .................... | .................... | .................... |
| F | 2916 hCoV-19/USA/CA-IGI-0079/2020\|EPI_ISL_511833\|2020-0 | .................... | .................... | ..................... | .................... | .................... | .................... |
| F | 2917 hCoV-19/USA/CA-IGI-0080/2020\|EPI_ISL_511834\|2020-0 | .................... | .................... | ..................... | .................... | .................... | .................... |
| F | 2918 hCoV-19/USA/CA-IGI-0081/2020\|EPI_ISL_511835\|2020-0 | .................... | .................... | ..................... | .................... | .................... | .................... |
| F | 2919 hCoV-19/USA/CA-IGI-0083/2020\|EPI_ISL_511837\|2020-0 | .................... | .................... | ..................... | .................... | .................... | .................... |
| F | 2920 hCoV-19/USA/CA-IGI-0085/2020\|EPI_ISL_511839\|2020-0 | .................... | .................... | ..................... | .................... | .................... | .................... |
| F | 2921 hCoV-19/USA/CA-IGI-0086/2020\|EPI_ISL_511840\|2020-0 | .................... | .................... | ..................... | .................... | .................... | .................... |
| F | 2922 hCoV-19/USA/CA-IGI-0088/2020\|EPI_ISL_511842\|2020-0 | .................... | .................... | ..................... | .................... | .................... | .................... |
| F | 2923 hCoV-19/USA/CA-IGI-0089/2020\|EPI_ISL_511843\|2020-0 | .................... | .................... | ..................... | .................... | .................... | .................... |
| F | 2924 hCoV-19/USA/CA-IGI-0091/2020\|EPI_ISL_511845\|2020-0 | .................... | .................... | ..................... | .................... | .................... | .................... |
| F | 2925 hCoV-19/USA/CA-IGI-0094/2020\|EPI_ISL_511848\|2020-0 | .................... | .................... | ..................... | .................... | .................... | .................... |
| F | 2926 hCoV-19/USA/CA-IGI-0095/2020\|EPI_ISL_511849\|2020-0 | .................... | .................... | ..................... | .................... | .................... | .................... |
| F | 2927 hCoV-19/USA/CA-IGI-0096/2020\|EPI_ISL_511850\|2020-0 | .................... | .................... | ..................... | .................... | .................... | .................... |
| F | 2928 hCoV-19/USA/CA-IGI-0097/2020\|EPI_ISL_511851\|2020-0 | .................... | .................... | ..................... | .................... | .................... | .................... |
| F | 2929 hCoV-19/USA/WA-UW-10034/2020\|EPI_ISL_511853\|2020-0 | .................... | .................... | ..................... | .................... | .................... | .................... |
| F | 2930 hCoV-19/USA/NV-NSPHL-A0013/2020\|EPI_ISL_515300\|202 | .................... | .................... | ..................... | .................... | .................... | .................... |
| F | 2931 hCoV-19/USA/NV-NSPHL-A0014/2020\|EPI_ISL_515301\|202 | .................... | .................... | ..................... | .................... | .................... | .................... |
| F | 2932 hCoV-19/USA/NV-NSPHL-A0015/2020\|EPI_ISL_515302\|202 | .................... | .................... | ..................... | .................... | .................... | .................... |
| F | 2933 hCoV-19/USA/NV-NSPHL-A0016/2020\|EPI_ISL_515303\|202 | .................... | .................... | ..................... | .................... | .................... | .................... |
| F | 2934 hCoV-19/USA/NV-NSPHL-A0017/2020\|EPI_ISL_515304\|202 | .................... | .................... | ..................... | .................... | .................... | .................... |
| F | 2935 hCoV-19/USA/NV-NSPHL-A0018/2020\|EPI_ISL_515305\|202 | .................... | .................... | ..................... | .................... | .................... | .................... |
| F | 2936 hCoV-19/USA/NV-NSPHL-A0019/2020\|EPI_ISL_515306\|202 | .................... | .................... | ..................... | .................... | .................... | .................... |
| F | 2937 hCoV-19/USA/NV-NSPHL-A0020/2020\|EPI_ISL_515307\|202 | .................... | .................... | ..................... | .................... | .................... | .................... |
| F | 2938 hCoV-19/USA/NV-NSPHL-A0021/2020\|EPI_ISL_515308\|202 | .................... | .................... | ..................... | .................... | .................... | .................... |
| F | 2939 hCoV-19/USA/NV-NSPHL-A0022/2020\|EPI_ISL_515309\|202 | .................... | .................... | ..................... | .................... | .................... | .................... |
| F | 2940 hCoV-19/USA/NV-NSPHL-A0024/2020\|EPI_ISL_515311\|202 | .................... | .................... | ..................... | .................... | .................... | .................... |
| F | 2941 hCoV-19/USA/NV-NSPHL-A0025/2020\|EPI_ISL_515312\|202 | .................... | .................... | ..................... | .................... | .................... | .................... |
| F | 2942 hCoV-19/USA/NV-NSPHL-A0026/2020\|EPI_ISL_515313\|202 | .................... | .................... | ..................... | .................... | .................... | .................... |
| F | 2943 hCoV-19/USA/NV-NSPHL-A0027/2020\|EPI_ISL_515314\|202 | .................... | .................... | ..................... | .................... | .................... | .................... |
| F | 2944 hCoV-19/USA/NV-NSPHL-A0028/2020\|EPI_ISL_515315\|202 | .................... | .................... | ..................... | .................... | .................... | .................... |
| F | 2945 hCoV-19/USA/NV-NSPHL-A0030/2020\|EPI_ISL_515316\|202 | .................... | .................... | ..................... | .................... | .................... | .................... |
| F | 2946 hCoV-19/USA/NV-NSPHL-A0031/2020\|EPI_ISL_515317\|202 | .................... | .................... | ..................... | .................... | .................... | .................... |
| F | 2947 hCoV-19/USA/NV-NSPHL-A0032/2020\|EPI_ISL_515318\|202 | .................... | .................... | ..................... | .................... | .................... | .................... |
| F | 2948 hCoV-19/USA/NV-NSPHL-A0033/2020\|EPI_ISL_515319\|202 | .................... | .................... | ..................... | .................... | .................... | .................... |
| F | 2949 hCoV-19/USA/NV-NSPHL-A0034/2020\|EPI_ISL_515320\|202 | .................... | .................... | ..................... | .................... | .................... | .................... |
| F | 2950 hCoV-19/USA/NV-NSPHL-A0036/2020\|EPI_ISL_515321\|202 | .................... | .................... | ..................... | .................... | .................... | .................... |
| F | 2951 hCoV-19/USA/NV-NSPHL-A0039/2020\|EPI_ISL_515322\|202 | .................... | .................... | ..................... | .................... | .................... | .................... |
| F | 2952 hCoV-19/USA/NV-NSPHL-A0041/2020\|EPI_ISL_515324\|202 | .................... | .................... | ..................... | .................... | .................... | .................... |
| F | 2953 hCoV-19/USA/NV-NSPHL-A0042/2020\|EPI_ISL_515325\|202 | .................... | .................... | ..................... | .................... | .................... | .................... |
| F | 2954 hCoV-19/USA/NV-NSPHL-A0044/2020\|EPI_ISL_515327\|202 | .................... | .................... | ..................... | .................... | .................... | .................... |
| F | 2955 hCoV-19/USA/NV-NSPHL-A0045/2020\|EPI_ISL_515328\|202 | .................... | .................... | ..................... | .................... | .................... | .................... |
| F | 2956 hCoV-19/USA/NV-NSPHL-A0046/2020\|EPI_ISL_515329\|202 | .................... | .................... | ..................... | .................... | .................... | .................... |
| F | 2957 hCoV-19/USA/NV-NSPHL-A0047/2020\|EPI_ISL_515330\|202 | .................... | .................... | ..................... | .................... | .................... | .................... |
| F | 2958 hCoV-19/USA/NV-NSPHL-A0048/2020\|EPI_ISL_515331\|202 | .................... | .................... | ..................... | .................... | .................... | .................... |
| F | 2959 hCoV-19/USA/NV-NSPHL-A0049/2020\|EPI_ISL_515332\|202 | .................... | .................... | ..................... | .................... | .................... | .................... |
| F | 2960 hCoV-19/USA/NV-NSPHL-A0051/2020\|EPI_ISL_515333\|202 | .................... | .................... | ..................... | .................... | .................... | .................... |
| F | 2961 hCoV-19/USA/NV-NSPHL-A0052/2020\|EPI_ISL_515334\|202 | .................... | .................... | ..................... | .................... | .................... | .................... |
| F | 2962 hCoV-19/USA/NV-NSPHL-A0054/2020\|EPI_ISL_515336\|202 | .................... | .................... | ..................... | .................... | .................... | .................... |
| F | 2963 hCoV-19/USA/NV-NSPHL-A0055/2020\|EPI_ISL_515337\|202 | .................... | .................... | ..................... | .................... | .................... | .................... |
| F | 2964 hCoV-19/USA/NV-NSPHL-A0056/2020\|EPI_ISL_515338\|202 | .................... | .................... | ..................... | .................... | .................... | .................... |
| F | 2965 hCoV-19/USA/NV-NSPHL-A0057/2020\|EPI_ISL_515339\|202 | .................... | .................... | ..................... | .................... | .................... | .................... |
| F | 2966 hCoV-19/USA/NV-NSPHL-A0058/2020\|EPI_ISL_515340\|202 | .................... | .................... | ..................... | .................... | .................... | .................... |
| F | 2967 hCoV-19/USA/NV-NSPHL-A0059/2020\|EPI_ISL_515341\|202 | .................... | .................... | ..................... | .................... | .................... | .................... |
| F | 2968 hCoV-19/USA/NV-NSPHL-A0060/2020\|EPI_ISL_515342\|202 | .................... | .................... | ..................... | .................... | .................... | .................... |
| F | 2969 hCoV-19/USA/NV-NSPHL-A0062/2020\|EPI_ISL_515344\|202 | .................... | .................... | ..................... | .................... | .................... | .................... |
| F | 2970 hCoV-19/USA/NV-NSPHL-A0064/2020\|EPI_ISL_515345\|202 | .................... | .................... | ..................... | .................... | .................... | .................... |
| F | 2971 hCoV-19/USA/NV-NSPHL-A0065/2020\|EPI_ISL_515346\|202 | .................... | .................... | ..................... | .................... | .................... | .................... |
| F | 2972 hCoV-19/USA/NV-NSPHL-A0066/2020\|EPI_ISL_515347\|202 | .................... | .................... | ..................... | .................... | .................... | .................... |
| F | 2973 hCoV-19/USA/NV-NSPHL-A0068/2020\|EPI_ISL_515348\|202 | .................... | .................... | ..................... | .................... | .................... | .................... |
| F | 2974 hCoV-19/USA/NV-NSPHL-A0069/2020\|EPI_ISL_515349\|202 | .................... | .................... | ..................... | .................... | .................... | .................... |
| F | 2975 hCoV-19/USA/NV-NSPHL-A0070/2020\|EPI_ISL_515350\|202 | .................... | .................... | ..................... | .................... | .................... | .................... |
| F | 2976 hCoV-19/USA/NV-NSPHL-A0071/2020\|EPI_ISL_515351\|202 | .................... | .................... | ..................... | .................... | .................... | .................... |
| F | 2977 hCoV-19/USA/NV-NSPHL-A0073/2020\|EPI_ISL_515352\|202 | .................... | .................... | ..................... | .................... | .................... | .................... |
| F | 2978 hCoV-19/USA/NV-NSPHL-A0075/2020\|EPI_ISL_515353\|202 | .................... | .................... | ..................... | .................... | .................... | .................... |
| F | 2979 hCoV-19/USA/NV-NSPHL-A0076/2020\|EPI_ISL_515354\|202 | .................... | .................... | ..................... | .................... | .................... | .................... |
| F | 2980 hCoV-19/USA/NV-NSPHL-A0078/2020\|EPI_ISL_515355\|202 | .................... | .................... | ..................... | .................... | .................... | .................... |
| F | 2981 hCoV-19/USA/NV-NSPHL-A0079/2020\|EPI_ISL_515356\|202 | .................... | .................... | ..................... | .................... | .................... | .................... |
| F | 2982 hCoV-19/USA/NV-NSPHL-A0080/2020\|EPI_ISL_515357\|202 | .................... | .................... | ..................... | .................... | .................... | .................... |
| F | 2983 hCoV-19/USA/NV-NSPHL-A0081/2020\|EPI_ISL_515358\|202 | .................... | .................... | ..................... | .................... | .................... | .................... |
| F | 2984 hCoV-19/USA/NV-NSPHL-A0082/2020\|EPI_ISL_515359\|202 | .................... | .................... | ..................... | .................... | .................... | .................... |
| F | 2985 hCoV-19/USA/NV-NSPHL-A0084/2020\|EPI_ISL_515361\|202 | .................... | .................... | ..................... | .................... | .................... | .................... |
| F | 2986 hCoV-19/USA/NV-NSPHL-A0085/2020\|EPI_ISL_515362\|202 | .................... | .................... | ..................... | .................... | .................... | .................... |
| F | 2987 hCoV-19/USA/NV-NSPHL-A0086/2020\|EPI_ISL_515363\|202 | .................... | .................... | ..................... | .................... | .................... | .................... |
| F | 2988 hCoV-19/USA/NV-NSPHL-A0087/2020\|EPI_ISL_515364\|202 | .................... | .................... | ..................... | .................... | .................... | .................... |
| F | 2989 hCoV-19/USA/NV-NSPHL-A0088/2020\|EPI_ISL_515365\|202 | .................... | .................... | ..................... | .................... | .................... | .................... |
| F | 2990 hCoV-19/USA/NV-NSPHL-A0089/2020\|EPI_ISL_515366\|202 | .................... | .................... | ..................... | .................... | .................... | .................... |
| F | 2991 hCoV-19/USA/NV-NSPHL-A0090/2020\|EPI_ISL_515367\|202 | .................... | .................... | ..................... | .................... | .................... | .................... |
| F | 2992 hCoV-19/USA/NV-NSPHL-A0091/2020\|EPI_ISL_515368\|202 | .................... | .................... | ..................... | .................... | .................... | .................... |

| | | | | | | | | |
|---|---|---|---|---|---|---|---|---|
| F | 2993 | hCoV-19/USA/NV-NSPHL-A0092/2020\|EPI_ISL_515369\|202 | .................... | .................... | .................... | .................... | .................... | .................... |
| F | 2994 | hCoV-19/USA/NV-NSPHL-A0093/2020\|EPI_ISL_515370\|202 | .................... | .................... | .................... | .................... | .................... | .................... |
| F | 2995 | hCoV-19/USA/NV-NSPHL-A0094/2020\|EPI_ISL_515371\|202 | .................... | .................... | .................... | .................... | .................... | .................... |
| F | 2996 | hCoV-19/USA/NV-NSPHL-A0095/2020\|EPI_ISL_515372\|202 | .................... | .................... | .................... | .................... | .................... | .................... |
| F | 2997 | hCoV-19/USA/NV-NSPHL-A0097/2020\|EPI_ISL_515373\|202 | .................... | .................... | .................... | .................... | .................... | .................... |
| F | 2998 | hCoV-19/USA/NV-NSPHL-A0098/2020\|EPI_ISL_515374\|202 | .................... | .................... | .................... | .................... | .................... | .................... |
| F | 2999 | hCoV-19/USA/NV-NSPHL-A0099/2020\|EPI_ISL_515375\|202 | .................... | .................... | .................... | .................... | .................... | .................... |
| F | 3000 | hCoV-19/USA/NV-NSPHL-A0103/2020\|EPI_ISL_515376\|202 | .................... | .................... | .................... | .................... | .................... | .................... |
| F | 3001 | hCoV-19/USA/NV-NSPHL-A0104/2020\|EPI_ISL_515377\|202 | .................... | .................... | .................... | .................... | .................... | .................... |
| F | 3002 | hCoV-19/USA/NV-NSPHL-A0105/2020\|EPI_ISL_515378\|202 | .................... | .................... | .................... | .................... | .................... | .................... |
| F | 3003 | hCoV-19/USA/NV-NSPHL-A0106/2020\|EPI_ISL_515379\|202 | .................... | .................... | .................... | .................... | .................... | .................... |
| F | 3004 | hCoV-19/Peru/ANC-INS-045/2020\|EPI_ISL_516628\|2020- | .................... | .................... | .................... | .................... | .................... | .................... |
| F | 3005 | hCoV-19/Germany/BE-ChVir-D712-1/2020\|EPI_ISL_51662 | .................... | .................... | .................... | .................... | .................... | .................... |
| F | 3006 | hCoV-19/Germany/BE-ChVir-D666-2/2020\|EPI_ISL_51663 | .................... | .................... | .................... | .................... | .................... | .................... |
| F | 3007 | hCoV-19/Germany/BE-ChVir-D718-3/2020\|EPI_ISL_51663 | .................... | .................... | .................... | .................... | .................... | .................... |
| F | 3008 | hCoV-19/Germany/BE-ChVir-D672-4/2020\|EPI_ISL_51663 | .................... | .................... | .................... | .................... | .................... | .................... |
| F | 3009 | hCoV-19/Germany/BE-ChVir-D658-5/2020\|EPI_ISL_51663 | .................... | .................... | .................... | .................... | .................... | .................... |
| F | 3010 | hCoV-19/Germany/BE-ChVir-D665-6/2020\|EPI_ISL_51663 | .................... | .................... | .................... | .................... | .................... | .................... |
| F | 3011 | hCoV-19/Germany/BE-ChVir-D667-7/2020\|EPI_ISL_51663 | .................... | .................... | .................... | .................... | .................... | .................... |
| F | 3012 | hCoV-19/Germany/BE-ChVir-D671-8/2020\|EPI_ISL_51663 | .................... | .................... | .................... | .................... | .................... | .................... |
| F | 3013 | hCoV-19/Germany/BE-ChVir-D670-9/2020\|EPI_ISL_51663 | .................... | .................... | .................... | .................... | .................... | .................... |
| F | 3014 | hCoV-19/Germany/BE-ChVir-D710-10/2020\|EPI_ISL_5166 | .................... | .................... | .................... | .................... | .................... | .................... |
| F | 3015 | hCoV-19/Germany/BE-ChVir-D711-11/2020\|EPI_ISL_5166 | .................... | .................... | .................... | .................... | .................... | .................... |
| F | 3016 | hCoV-19/Germany/BE-ChVir-D717-D761-12/2020\|EPI_ISL | .................... | .................... | .................... | .................... | .................... | .................... |
| F | 3017 | hCoV-19/Germany/BE-ChVir-W1191-13/2020\|EPI_ISL_516 | .................... | .................... | .................... | .................... | .................... | .................... |
| F | 3018 | hCoV-19/Germany/BE-ChVir-D679-14/2020\|EPI_ISL_5166 | .................... | .................... | .................... | .................... | .................... | .................... |
| F | 3019 | hCoV-19/Germany/BE-ChVir-W1248-16/2020\|EPI_ISL_516 | .................... | .................... | .................... | .................... | .................... | .................... |
| F | 3020 | hCoV-19/Germany/BE-ChVir-D715-D799-17/2020\|EPI_ISL | .................... | .................... | .................... | .................... | .................... | .................... |
| F | 3021 | hCoV-19/Peru/LIM-INS-046/2020\|EPI_ISL_516646\|2020- | .................... | .................... | .................... | .................... | .................... | .................... |
| F | 3022 | hCoV-19/Peru/JUN-INS-047/2020\|EPI_ISL_516647\|2020- | .................... | .................... | .................... | .................... | .................... | .................... |
| F | 3023 | hCoV-19/Ecuador/USFQ-161/2020\|EPI_ISL_516648\|2020- | .................... | .................... | .................... | .................... | .................... | .................... |
| F | 3024 | hCoV-19/Ecuador/USFQ-162/2020\|EPI_ISL_516649\|2020- | .................... | .................... | .................... | .................... | .................... | .................... |
| F | 3025 | hCoV-19/Ecuador/USFQ-165/2020\|EPI_ISL_516650\|2020- | .................... | .................... | .................... | .................... | .................... | .................... |
| F | 3026 | hCoV-19/Ecuador/USFQ-167/2020\|EPI_ISL_516651\|2020- | .................... | .................... | .................... | .................... | .................... | .................... |
| F | 3027 | hCoV-19/Ecuador/USFQ-171/2020\|EPI_ISL_516652\|2020- | .................... | .................... | .................... | .................... | .................... | .................... |
| F | 3028 | hCoV-19/Peru/ANC-INS-049/2020\|EPI_ISL_516653\|2020- | .................... | .................... | .................... | .................... | .................... | .................... |
| F | 3029 | hCoV-19/Peru/ANC-INS-050/2020\|EPI_ISL_516654\|2020- | .................... | .................... | .................... | .................... | .................... | .................... |
| F | 3030 | hCoV-19/USA/VA-DCLS-0859/2020\|EPI_ISL_516655\|2020- | .................... | .................... | .................... | .................... | .................... | .................... |
| F | 3031 | hCoV-19/USA/VA-DCLS-0860/2020\|EPI_ISL_516656\|2020- | .................... | .................... | .................... | .................... | .................... | .................... |
| F | 3032 | hCoV-19/USA/VA-DCLS-0861/2020\|EPI_ISL_516657\|2020- | .................... | .................... | .................... | .................... | .................... | .................... |
| F | 3033 | hCoV-19/USA/VA-DCLS-0863/2020\|EPI_ISL_516659\|2020- | .................... | .................... | .................... | .................... | .................... | .................... |
| F | 3034 | hCoV-19/USA/VA-DCLS-0864/2020\|EPI_ISL_516660\|2020- | .................... | .................... | .................... | .................... | .................... | .................... |
| F | 3035 | hCoV-19/USA/VA-DCLS-0865/2020\|EPI_ISL_516661\|2020- | .................... | .................... | .................... | .................... | .................... | .................... |
| F | 3036 | hCoV-19/USA/VA-DCLS-0867/2020\|EPI_ISL_516662\|2020- | .................... | .................... | .................... | .................... | .................... | .................... |
| F | 3037 | hCoV-19/USA/VA-DCLS-0868/2020\|EPI_ISL_516663\|2020- | .................... | .................... | .................... | .................... | .................... | .................... |
| F | 3038 | hCoV-19/USA/VA-DCLS-0869/2020\|EPI_ISL_516664\|2020- | .................... | .................... | .................... | .................... | .................... | .................... |
| F | 3039 | hCoV-19/USA/VA-DCLS-0870/2020\|EPI_ISL_516665\|2020- | .................... | .................... | .................... | .................... | .................... | .................... |
| F | 3040 | hCoV-19/USA/VA-DCLS-0871/2020\|EPI_ISL_516666\|2020- | .................... | .................... | .................... | .................... | .................... | .................... |
| F | 3041 | hCoV-19/USA/VA-DCLS-0876/2020\|EPI_ISL_516667\|2020- | .................... | .................... | .................... | .................... | .................... | .................... |
| F | 3042 | hCoV-19/USA/VA-DCLS-0877/2020\|EPI_ISL_516668\|2020- | .................... | .................... | .................... | .................... | .................... | .................... |
| F | 3043 | hCoV-19/USA/VA-DCLS-0878/2020\|EPI_ISL_516669\|2020- | .................... | .................... | .................... | .................... | .................... | .................... |
| F | 3044 | hCoV-19/USA/VA-DCLS-0880/2020\|EPI_ISL_516670\|2020- | .................... | .................... | .................... | .................... | .................... | .................... |
| F | 3045 | hCoV-19/USA/VA-DCLS-0881/2020\|EPI_ISL_516671\|2020- | .................... | .................... | .................... | .................... | .................... | .................... |
| F | 3046 | hCoV-19/USA/VA-DCLS-0882/2020\|EPI_ISL_516672\|2020- | .................... | .................... | .................... | .................... | .................... | .................... |
| F | 3047 | hCoV-19/USA/VA-DCLS-0883/2020\|EPI_ISL_516673\|2020- | .................... | .................... | .................... | .................... | .................... | .................... |
| F | 3048 | hCoV-19/USA/VA-DCLS-0884/2020\|EPI_ISL_516674\|2020- | .................... | .................... | .................... | .................... | .................... | .................... |
| F | 3050 | hCoV-19/USA/VA-DCLS-0887/2020\|EPI_ISL_516676\|2020- | .................... | .................... | .................... | .................... | .................... | .................... |
| F | 3051 | hCoV-19/USA/VA-DCLS-0888/2020\|EPI_ISL_516677\|2020- | .................... | .................... | .................... | .................... | .................... | .................... |
| F | 3052 | hCoV-19/USA/VA-DCLS-0891/2020\|EPI_ISL_516678\|2020- | .................... | .................... | .................... | .................... | .................... | .................... |
| F | 3053 | hCoV-19/USA/VA-DCLS-0893/2020\|EPI_ISL_516679\|2020- | .................... | .................... | .................... | .................... | .................... | .................... |
| F | 3054 | hCoV-19/USA/VA-DCLS-0894/2020\|EPI_ISL_516680\|2020- | .................... | .................... | .................... | .................... | .................... | .................... |
| F | 3055 | hCoV-19/USA/VA-DCLS-0896/2020\|EPI_ISL_516681\|2020- | .................... | .................... | .................... | .................... | .................... | .................... |
| F | 3056 | hCoV-19/USA/VA-DCLS-0897/2020\|EPI_ISL_516682\|2020- | .................... | .................... | .................... | .................... | .................... | .................... |
| F | 3057 | hCoV-19/USA/VA-DCLS-0898/2020\|EPI_ISL_516683\|2020- | .................... | .................... | .................... | .................... | .................... | .................... |
| F | 3058 | hCoV-19/USA/VA-DCLS-0899/2020\|EPI_ISL_516684\|2020- | .................... | .................... | .................... | .................... | .................... | .................... |
| F | 3059 | hCoV-19/Peru/ANC-INS-051/2020\|EPI_ISL_516685\|2020- | .................... | .................... | .................... | .................... | .................... | .................... |
| F | 3060 | hCoV-19/Peru/ANC-INS-052/2020\|EPI_ISL_516686\|2020- | .................... | .................... | .................... | .................... | .................... | .................... |
| F | 3061 | hCoV-19/USA/VA-DCLS-0358/2020\|EPI_ISL_516687\|2020- | .................... | .................... | .................... | .................... | .................... | .................... |
| F | 3062 | hCoV-19/USA/VA-DCLS-0359/2020\|EPI_ISL_516688\|2020- | .................... | .................... | .................... | .................... | .................... | .................... |
| F | 3063 | hCoV-19/USA/VA-DCLS-0360/2020\|EPI_ISL_516689\|2020- | .................... | .................... | .................... | .................... | .................... | .................... |
| F | 3064 | hCoV-19/USA/VA-DCLS-0361/2020\|EPI_ISL_516690\|2020- | .................... | .................... | .................... | .................... | .................... | .................... |
| F | 3065 | hCoV-19/USA/VA-DCLS-0362/2020\|EPI_ISL_516691\|2020- | .................... | .................... | .................... | .................... | .................... | .................... |
| F | 3066 | hCoV-19/USA/VA-DCLS-0369/2020\|EPI_ISL_516692\|2020- | .................... | .................... | .................... | .................... | .................... | .................... |
| F | 3067 | hCoV-19/USA/VA-DCLS-0370/2020\|EPI_ISL_516693\|2020- | .................... | .................... | .................... | .................... | .................... | .................... |
| F | 3068 | hCoV-19/USA/VA-DCLS-0813/2020\|EPI_ISL_516694\|2020- | .................... | .................... | .................... | .................... | .................... | .................... |
| F | 3069 | hCoV-19/USA/VA-DCLS-0816/2020\|EPI_ISL_516695\|2020- | .................... | .................... | .................... | .................... | .................... | .................... |
| F | 3070 | hCoV-19/USA/VA-DCLS-0818/2020\|EPI_ISL_516696\|2020- | .................... | .................... | .................... | .................... | .................... | .................... |
| F | 3071 | hCoV-19/USA/VA-DCLS-0824/2020\|EPI_ISL_516698\|2020- | .................... | .................... | .................... | .................... | .................... | .................... |
| F | 3072 | hCoV-19/USA/VA-DCLS-0825/2020\|EPI_ISL_516699\|2020- | .................... | .................... | .................... | .................... | .................... | .................... |
| F | 3073 | hCoV-19/USA/WA-S1885/2020\|EPI_ISL_496311\|2020-05-2 | .................... | .................... | .................... | .................... | .................... | .................... |

| | | | | | | | | |
|---|---|---|---|---|---|---|---|---|
| F | 3074 hCoV-19/USA/WA-S1886/2020\|EPI_ISL_496312\|2020-05-2 | ···················· | ···················· | ···················· | ···················· | ···················· | ···················· | |
| F | 3075 hCoV-19/USA/WA-S1887/2020\|EPI_ISL_496313\|2020-05-2 | ···················· | ···················· | ···················· | ···················· | ···················· | ···················· | |
| F | 3076 hCoV-19/USA/WA-S1888/2020\|EPI_ISL_496314\|2020-05-2 | ···················· | ···················· | ···················· | ···················· | ···················· | ···················· | |
| F | 3077 hCoV-19/USA/WA-S1896/2020\|EPI_ISL_496322\|2020-05-2 | ···················· | ···················· | ···················· | ···················· | ···················· | ···················· | |
| F | 3078 hCoV-19/USA/WA-S1897/2020\|EPI_ISL_496323\|2020-05-2 | ···················· | ···················· | ···················· | ···················· | ···················· | ···················· | |
| F | 3079 hCoV-19/USA/WA-S1898/2020\|EPI_ISL_496324\|2020-05-2 | ···················· | ···················· | ···················· | ···················· | ···················· | ···················· | |
| F | 3080 hCoV-19/USA/WA-S1899/2020\|EPI_ISL_496325\|2020-05-2 | ···················· | ···················· | ···················· | ···················· | ···················· | ···················· | |
| F | 3081 hCoV-19/USA/WA-S1900/2020\|EPI_ISL_496326\|2020-05-2 | ···················· | ···················· | ···················· | ···················· | ···················· | ···················· | |
| F | 3082 hCoV-19/USA/WA-S1901/2020\|EPI_ISL_496327\|2020-05-2 | ···················· | ···················· | ···················· | ···················· | ···················· | ···················· | |
| F | 3083 hCoV-19/USA/WA-S1902/2020\|EPI_ISL_496328\|2020-05-2 | ···················· | ···················· | ···················· | ···················· | ···················· | ···················· | |
| F | 3084 hCoV-19/USA/WA-S1903/2020\|EPI_ISL_496329\|2020-05-2 | ···················· | ···················· | ···················· | ···················· | ···················· | ···················· | |
| F | 3085 hCoV-19/USA/WA-S1904/2020\|EPI_ISL_496330\|2020-05-2 | ···················· | ···················· | ···················· | ···················· | ···················· | ···················· | |
| F | 3086 hCoV-19/USA/WA-S1905/2020\|EPI_ISL_496331\|2020-05-2 | ···················· | ···················· | ···················· | ···················· | ···················· | ···················· | |
| F | 3087 hCoV-19/USA/WA-S1906/2020\|EPI_ISL_496332\|2020-05-2 | ···················· | ···················· | ···················· | ···················· | ···················· | ···················· | |
| F | 3088 hCoV-19/Mexico/BCN-ALSR-1708/2020\|EPI_ISL_496369\|2 | ···················· | ·A·················· | ···················· | ···················· | ···················· | ···················· | |
| F | 3089 hCoV-19/Mexico/BCN-ALSR-1713/2020\|EPI_ISL_496374\|2 | ···················· | ·A·················· | ···················· | ···················· | ···················· | ···················· | |
| F | 3090 hCoV-19/USA/WA-S2458/2020\|EPI_ISL_497090\|2020-06-1 | ···················· | ···················· | ···················· | ···················· | ···················· | ···················· | |
| F | 3091 hCoV-19/USA/WA-S2459/2020\|EPI_ISL_497091\|2020-06-1 | ···················· | ···················· | ···················· | ···················· | ···················· | ···················· | |
| F | 3092 hCoV-19/USA/WA-S2460/2020\|EPI_ISL_497092\|2020-06-1 | ···················· | ···················· | ···················· | ···················· | ···················· | ···················· | |
| F | 3093 hCoV-19/USA/WA-S2461/2020\|EPI_ISL_497093\|2020-06-1 | ···················· | ···················· | ···················· | ···················· | ···················· | ···················· | |
| F | 3094 hCoV-19/USA/WA-S2463/2020\|EPI_ISL_497095\|2020-06-1 | ···················· | ···················· | ···················· | ···················· | ···················· | ···················· | |
| F | 3095 hCoV-19/USA/WA-S2464/2020\|EPI_ISL_497096\|2020-06-1 | ···················· | ···················· | ···················· | ···················· | ···················· | ···················· | |
| F | 3096 hCoV-19/USA/WA-S2465/2020\|EPI_ISL_497097\|2020-06-1 | ···················· | ···················· | ···················· | ···················· | ···················· | ···················· | |
| F | 3097 hCoV-19/USA/WA-S2466/2020\|EPI_ISL_497098\|2020-06-1 | ···················· | ···················· | ···················· | ···················· | ···················· | ···················· | |
| F | 3098 hCoV-19/USA/WA-S2467/2020\|EPI_ISL_497099\|2020-06-1 | ···················· | ···················· | ···················· | ···················· | ···················· | ···················· | |
| F | 3099 hCoV-19/USA/WA-S2214/2020\|EPI_ISL_497642\|2020-06-0 | ···················· | ···················· | ···················· | ···················· | ···················· | ···················· | |
| F | 3100 hCoV-19/USA/WA-S2215/2020\|EPI_ISL_497643\|2020-06-0 | ···················· | ···················· | ···················· | ···················· | ···················· | ···················· | |
| F | 3101 hCoV-19/USA/WA-S2216/2020\|EPI_ISL_497644\|2020-06-0 | ···················· | ···················· | ···················· | ···················· | ···················· | ···················· | |
| F | 3102 hCoV-19/USA/WA-S2217/2020\|EPI_ISL_497645\|2020-06-0 | ···················· | ···················· | ···················· | ···················· | ···················· | ···················· | |
| F | 3103 hCoV-19/USA/WA-S2221/2020\|EPI_ISL_497649\|2020-06-0 | ···················· | ···················· | ···················· | ···················· | ···················· | ···················· | |
| F | 3104 hCoV-19/USA/WA-S2226/2020\|EPI_ISL_497654\|2020-06-0 | ···················· | ···················· | ···················· | ···················· | ···················· | ···················· | |
| F | 3105 hCoV-19/USA/WA-S2227/2020\|EPI_ISL_497655\|2020-06-0 | ···················· | ···················· | ···················· | ···················· | ···················· | ···················· | |
| F | 3106 hCoV-19/USA/WA-S2228/2020\|EPI_ISL_497656\|2020-06-0 | ···················· | ···················· | ···················· | ···················· | ···················· | ···················· | |
| F | 3107 hCoV-19/USA/WA-S2229/2020\|EPI_ISL_497657\|2020-06-0 | ···················· | ···················· | ···················· | ···················· | ···················· | ···················· | |
| F | 3108 hCoV-19/USA/WA-S2230/2020\|EPI_ISL_497658\|2020-06-1 | ···················· | ···················· | ···················· | ···················· | ···················· | ···················· | |
| F | 3109 hCoV-19/USA/WA-S2231/2020\|EPI_ISL_497659\|2020-06-0 | ···················· | ···················· | ···················· | ···················· | ···················· | ···················· | |
| F | 3110 hCoV-19/USA/WA-S2232/2020\|EPI_ISL_497660\|2020-06-0 | ···················· | ···················· | ···················· | ···················· | ···················· | ···················· | |
| F | 3111 hCoV-19/USA/WA-S2233/2020\|EPI_ISL_497661\|2020-06-0 | ···················· | ···················· | ···················· | ···················· | ···················· | ···················· | |
| F | 3112 hCoV-19/USA/WA-S2234/2020\|EPI_ISL_497662\|2020-06-0 | ···················· | ···················· | ···················· | ···················· | ···················· | ···················· | |
| F | 3113 hCoV-19/USA/WA-S2235/2020\|EPI_ISL_497663\|2020-06-0 | ···················· | ···················· | ···················· | ···················· | ···················· | ···················· | |
| F | 3114 hCoV-19/USA/WA-S2258/2020\|EPI_ISL_497686\|2020-06-0 | ···················· | ···················· | ···················· | ···················· | ···················· | ···················· | |
| F | 3115 hCoV-19/USA/WA-S2259/2020\|EPI_ISL_497687\|2020-06-0 | ···················· | ···················· | ···················· | ···················· | ···················· | ···················· | |
| F | 3116 hCoV-19/USA/WA-S2260/2020\|EPI_ISL_497688\|2020-06-0 | ···················· | ···················· | ···················· | ···················· | ···················· | ···················· | |
| F | 3117 hCoV-19/USA/WA-S2261/2020\|EPI_ISL_497689\|2020-06-0 | ···················· | ···················· | ···················· | ···················· | ···················· | ···················· | |
| F | 3118 hCoV-19/USA/WA-S2262/2020\|EPI_ISL_497690\|2020-06-1 | ···················· | ···················· | ···················· | ···················· | ···················· | ···················· | |
| F | 3119 hCoV-19/USA/WA-S2263/2020\|EPI_ISL_497691\|2020-06-0 | ···················· | ···················· | ···················· | ···················· | ···················· | ···················· | |
| F | 3120 hCoV-19/USA/WA-S2264/2020\|EPI_ISL_497692\|2020-06-0 | ···················· | ···················· | ···················· | ···················· | ···················· | ···················· | |
| F | 3121 hCoV-19/USA/WA-S2265/2020\|EPI_ISL_497693\|2020-06-0 | ···················· | ···················· | ···················· | ···················· | ···················· | ···················· | |
| F | 3122 hCoV-19/USA/WA-S2266/2020\|EPI_ISL_497694\|2020-06-0 | ···················· | ···················· | ···················· | ···················· | ···················· | ···················· | |
| F | 3123 hCoV-19/USA/WA-S2267/2020\|EPI_ISL_497695\|2020-06-0 | ···················· | ···················· | ···················· | ···················· | ···················· | ···················· | |
| F | 3124 hCoV-19/USA/WA-S2268/2020\|EPI_ISL_497696\|2020-06-0 | ···················· | ···················· | ···················· | ···················· | ···················· | ···················· | |
| F | 3125 hCoV-19/USA/WA-S2269/2020\|EPI_ISL_497697\|2020-06-0 | ···················· | ···················· | ···················· | ···················· | ···················· | ···················· | |
| F | 3126 hCoV-19/USA/WA-S2270/2020\|EPI_ISL_497698\|2020-06-0 | ···················· | ···················· | ···················· | ···················· | ···················· | ···················· | |
| F | 3127 hCoV-19/USA/WA-S2271/2020\|EPI_ISL_497699\|2020-06-1 | ···················· | ···················· | ···················· | ···················· | ···················· | ···················· | |
| F | 3128 hCoV-19/England/LIVE-A50D0/2020\|EPI_ISL_500106\|202 | ···················· | ···················· | ···················· | ···················· | ···················· | ···················· | |
| F | 3129 hCoV-19/England/LIVE-A50FE/2020\|EPI_ISL_500107\|202 | ···················· | ···················· | ···················· | ···················· | ···················· | ···················· | |
| F | 3130 hCoV-19/England/LIVE-A510A/2020\|EPI_ISL_500108\|202 | ···················· | ···················· | ···················· | ···················· | ···················· | ···················· | |
| F | 3131 hCoV-19/England/LIVE-A5155/2020\|EPI_ISL_500109\|202 | ···················· | ···················· | ···················· | ···················· | ···················· | ···················· | |
| F | 3132 hCoV-19/England/LIVE-A5191/2020\|EPI_ISL_500110\|202 | ···················· | ···················· | ···················· | ···················· | ···················· | ···················· | |
| F | 3134 hCoV-19/England/LIVE-A5207/2020\|EPI_ISL_500112\|202 | ···················· | ···················· | ···················· | ···················· | ···················· | ···················· | |
| F | 3135 hCoV-19/England/LIVE-A5252/2020\|EPI_ISL_500113\|202 | ···················· | ···················· | ···················· | ···················· | ···················· | ···················· | |
| F | 3137 hCoV-19/England/LIVE-A53D7/2020\|EPI_ISL_500115\|202 | ···················· | ···················· | ···················· | ···················· | ···················· | ···················· | |
| F | 3138 hCoV-19/England/LIVE-A545C/2020\|EPI_ISL_500116\|202 | ···················· | ···················· | ···················· | ···················· | ···················· | ···················· | |
| F | 3141 hCoV-19/England/LIVE-A55C2/2020\|EPI_ISL_500130\|202 | ···················· | ···················· | ···················· | ···················· | ···················· | ···················· | |
| F | 3142 hCoV-19/England/LIVE-A55D1/2020\|EPI_ISL_500131\|202 | ···················· | ···················· | ···················· | ···················· | ···················· | ···················· | |
| F | 3143 hCoV-19/England/LIVE-A55FF/2020\|EPI_ISL_500132\|202 | ···················· | ···················· | ···················· | ···················· | ···················· | ···················· | |
| F | 3144 hCoV-19/England/LIVE-A561A/2020\|EPI_ISL_500133\|202 | ···················· | ···················· | ···················· | ···················· | ···················· | ···················· | |
| F | 3145 hCoV-19/England/LIVE-A5629/2020\|EPI_ISL_500134\|202 | ···················· | ···················· | ···················· | ···················· | ···················· | ···················· | |
| F | 3146 hCoV-19/England/LIVE-A5647/2020\|EPI_ISL_500135\|202 | ···················· | ···················· | ···················· | ···················· | ···················N | ···················· | 1 |
| F | 3147 hCoV-19/England/LIVE-A5674/2020\|EPI_ISL_500136\|202 | ···················· | ···················· | ···················· | ···················· | ···················· | ···················· | |
| F | 3150 hCoV-19/England/LIVE-A56B0/2020\|EPI_ISL_500139\|202 | ···················· | ···················· | ···················· | ···················· | ···················· | ···················· | |
| F | 3151 hCoV-19/England/LIVE-A56CF/2020\|EPI_ISL_500140\|202 | ···················· | ···················· | ···················· | ···················· | ···················· | ···················· | |
| F | 3158 hCoV-19/England/LIVE-A5762/2020\|EPI_ISL_500147\|202 | ···················· | ···················· | ···················· | ···················· | ···················· | ···················· | |
| F | 3159 hCoV-19/England/LIVE-A5780/2020\|EPI_ISL_500148\|202 | ···················· | ···················· | ···················· | ···················· | ···················· | ···················· | |
| F | 3161 hCoV-19/England/LIVE-A57AE/2020\|EPI_ISL_500150\|202 | ···················· | ···················· | ···················· | ···················· | ···················· | ···················· | |
| F | 3163 hCoV-19/England/LIVE-A57DB/2020\|EPI_ISL_500152\|202 | ···················· | ···················· | ···················· | ···················· | ···················· | ···················· | |
| F | 3164 hCoV-19/England/LIVE-A57EA/2020\|EPI_ISL_500153\|202 | ···················· | ···················· | ···················· | ···················· | ···················· | ···················· | |
| F | 3166 hCoV-19/England/LIVE-A5814/2020\|EPI_ISL_500155\|202 | ···················· | ···················· | ···················· | ···················· | ···················· | ···················· | |
| F | 3168 hCoV-19/Australia/NSW2184/2020\|EPI_ISL_509510\|2020 | ···················· | ···················· | ···················· | ···················· | ···················· | ···················· | |
| F | 3169 hCoV-19/Australia/NSW2232/2020\|EPI_ISL_509511\|2020 | ···················· | ···················· | ···················· | ···················· | ···················· | ···················· | |

| | | | | | | | | |
|---|---|---|---|---|---|---|---|---|
| F | 3170 | hCoV-19/Australia/NSW2233/2020\|EPI_ISL_509512\|2020 | .................... | .................... | ..................... | .................... | .................... | .................... |
| F | 3171 | hCoV-19/Australia/NSW2234/2020\|EPI_ISL_509513\|2020 | .................... | .................... | ..................... | .................... | .................... | .................... |
| F | 3172 | hCoV-19/Australia/NSW2274/2020\|EPI_ISL_509518\|2020 | .................... | .................... | ..................... | .................... | .................... | .................... |
| F | 3173 | hCoV-19/Australia/NSW2280/2020\|EPI_ISL_509519\|2020 | .................... | .................... | ..................... | .................... | .................... | .................... |
| F | 3174 | hCoV-19/USA/CA-IGI-0050/2020\|EPI_ISL_511805\|2020-0 | .................... | .................... | ..................... | .................... | .................... | .................... |
| F | 3175 | hCoV-19/USA/CA-IGI-0051/2020\|EPI_ISL_511806\|2020-0 | .................... | .................... | ..................... | .................... | .................... | .................... |
| F | 3176 | hCoV-19/USA/CA-IGI-0052/2020\|EPI_ISL_511807\|2020-0 | .................... | .................... | ..................... | .................... | .................... | .................... |
| F | 3177 | hCoV-19/USA/CA-IGI-0053/2020\|EPI_ISL_511808\|2020-0 | .................... | .................... | ..................... | .................... | .................... | .................... |
| F | 3178 | hCoV-19/USA/CA-IGI-0054/2020\|EPI_ISL_511809\|2020-0 | .................... | .................... | ..................... | .................... | .................... | .................... |
| F | 3179 | hCoV-19/USA/CA-IGI-0058/2020\|EPI_ISL_511813\|2020-0 | .................... | .................... | ..................... | .................... | .................... | .................... |
| F | 3180 | hCoV-19/USA/CA-IGI-0064/2020\|EPI_ISL_511819\|2020-0 | .................... | .................... | ..................... | .................... | .................... | .................... |
| F | 3181 | hCoV-19/Ireland/CO-NVRL-71IRL82463/2020\|EPI_ISL_51 | .................... | .................... | ..................... | .................... | .................... | .................... |
| F | 3182 | hCoV-19/Ireland/D-NVRL-71IRL85733/2020\|EPI_ISL_512 | .................... | .................... | ..................... | .................... | .................... | .................... |
| F | 3183 | hCoV-19/Ireland/D-NVRL-71IRL86818/2020\|EPI_ISL_512 | .................... | .................... | ..................... | .................... | .................... | .................... |
| F | 3184 | hCoV-19/Ireland/D-NVRL-71IRL86828/2020\|EPI_ISL_512 | .................... | .................... | ..................... | .................... | .................... | .................... |
| F | 3185 | hCoV-19/Ireland/KE-NVRL-71IRL87566/2020\|EPI_ISL_51 | .................... | .................... | ..................... | .................... | .................... | .................... |
| F | 3186 | hCoV-19/Ireland/KE-NVRL-71IRL87592/2020\|EPI_ISL_51 | .................... | .................... | ..................... | .................... | .................... | .................... |
| F | 3187 | hCoV-19/USA/AK-PHL522/2020\|EPI_ISL_512144\|2020-06- | .................... | .................... | ..................... | .................... | .................... | .................... |
| F | 3188 | hCoV-19/USA/AK-PHL575/2020\|EPI_ISL_512148\|2020-06- | .................... | .................... | ..................... | .................... | .................... | .................... |
| F | 3189 | hCoV-19/USA/ME-HETL-J0014/2020\|EPI_ISL_513427\|2020 | .................... | .................... | ..................... | .................... | .................... | .................... |
| F | 3190 | hCoV-19/USA/ME-HETL-J0015/2020\|EPI_ISL_513428\|2020 | .................... | .................... | ..................... | .................... | .................... | .................... |
| F | 3191 | hCoV-19/USA/ME-HETL-J0016/2020\|EPI_ISL_513429\|2020 | .................... | .................... | ..................... | .................... | .................... | .................... |
| F | 3192 | hCoV-19/USA/ME-HETL-J0017/2020\|EPI_ISL_513430\|2020 | .................... | .................... | ..................... | .................... | .................... | .................... |
| F | 3193 | hCoV-19/USA/ME-HETL-J0018/2020\|EPI_ISL_513431\|2020 | .................... | .................... | ..................... | .................... | .................... | .................... |
| F | 3194 | hCoV-19/USA/ME-HETL-J0019/2020\|EPI_ISL_513432\|2020 | .................... | .................... | ..................... | .................... | .................... | .................... |
| F | 3195 | hCoV-19/USA/ME-HETL-J0020/2020\|EPI_ISL_513433\|2020 | .................... | .................... | ..................... | .................... | .................... | .................... |
| F | 3196 | hCoV-19/USA/ME-HETL-J0022/2020\|EPI_ISL_513434\|2020 | .................... | .................... | ..................... | .................... | .................... | .................... |
| F | 3197 | hCoV-19/USA/ME-HETL-J0023/2020\|EPI_ISL_513435\|2020 | .................... | .................... | ..................... | .................... | .................... | .................... |
| F | 3198 | hCoV-19/USA/ME-HETL-J0024/2020\|EPI_ISL_513436\|2020 | .................... | .................... | ..................... | .................... | .................... | .................... |
| F | 3199 | hCoV-19/USA/ME-HETL-J0025/2020\|EPI_ISL_513437\|2020 | .................... | .................... | ..................... | .................... | .................... | .................... |
| F | 3200 | hCoV-19/USA/ME-HETL-J0026/2020\|EPI_ISL_513438\|2020 | .................... | .................... | ..................... | .................... | .................... | .................... |
| F | 3201 | hCoV-19/USA/ME-HETL-J0027/2020\|EPI_ISL_513439\|2020 | .................... | .................... | ..................... | .................... | .................... | .................... |
| F | 3202 | hCoV-19/USA/ME-HETL-J0028/2020\|EPI_ISL_513440\|2020 | .................... | .................... | ..................... | .................... | .................... | .................... |
| F | 3203 | hCoV-19/USA/ME-HETL-J0029/2020\|EPI_ISL_513441\|2020 | .................... | .................... | ..................... | .................... | .................... | .................... |
| F | 3204 | hCoV-19/USA/ME-HETL-J0030/2020\|EPI_ISL_513442\|2020 | .................... | .................... | ..................... | .................... | .................... | .................... |
| F | 3205 | hCoV-19/USA/ME-HETL-J0031/2020\|EPI_ISL_513443\|2020 | .................... | .................... | ..................... | .................... | .................... | .................... |
| F | 3206 | hCoV-19/USA/ME-HETL-J0032/2020\|EPI_ISL_513444\|2020 | .................... | .................... | ..................... | .................... | .................... | .................... |
| F | 3207 | hCoV-19/USA/ME-HETL-J0035/2020\|EPI_ISL_513447\|2020 | .................... | .................... | ..................... | .................... | .................... | .................... |
| F | 3208 | hCoV-19/USA/ME-HETL-J0036/2020\|EPI_ISL_513448\|2020 | .................... | .................... | ..................... | .................... | .................... | .................... |
| F | 3209 | hCoV-19/USA/ME-HETL-J0037/2020\|EPI_ISL_513449\|2020 | .................... | .................... | ..................... | .................... | .................... | .................... |
| F | 3210 | hCoV-19/USA/ME-HETL-J0038/2020\|EPI_ISL_513450\|2020 | .................... | .................... | ..................... | .................... | .................... | .................... |
| F | 3211 | hCoV-19/USA/ME-HETL-J0039/2020\|EPI_ISL_513451\|2020 | .................... | .................... | ..................... | .................... | .................... | .................... |
| F | 3212 | hCoV-19/USA/ME-HETL-J0040/2020\|EPI_ISL_513452\|2020 | .................... | .................... | ..................... | .................... | .................... | .................... |
| F | 3213 | hCoV-19/USA/ME-HETL-J0041/2020\|EPI_ISL_513453\|2020 | .................... | .................... | ..................... | .................... | .................... | .................... |
| F | 3214 | hCoV-19/USA/ME-HETL-J0042/2020\|EPI_ISL_513454\|2020 | .................... | .................... | ..................... | .................... | .................... | .................... |
| F | 3215 | hCoV-19/USA/ME-HETL-J0043/2020\|EPI_ISL_513455\|2020 | .................... | .................... | ..................... | .................... | .................... | .................... |
| F | 3216 | hCoV-19/USA/ME-HETL-J0044/2020\|EPI_ISL_513456\|2020 | .................... | .................... | ..................... | .................... | .................... | .................... |
| F | 3217 | hCoV-19/USA/ME-HETL-J0046/2020\|EPI_ISL_513457\|2020 | .................... | .................... | ..................... | .................... | .................... | .................... |
| F | 3218 | hCoV-19/USA/ME-HETL-J0047/2020\|EPI_ISL_513458\|2020 | .................... | .................... | ..................... | .................... | .................... | .................... |
| F | 3219 | hCoV-19/USA/ME-HETL-J0048/2020\|EPI_ISL_513459\|2020 | .................... | .................... | ..................... | .................... | .................... | .................... |
| F | 3220 | hCoV-19/USA/ME-HETL-J0049/2020\|EPI_ISL_513460\|2020 | .................... | .................... | ..................... | .................... | .................... | .................... |
| F | 3221 | hCoV-19/USA/ME-HETL-J0050/2020\|EPI_ISL_513461\|2020 | .................... | .................... | ..................... | .................... | .................... | .................... |
| F | 3222 | hCoV-19/USA/ME-HETL-J0051/2020\|EPI_ISL_513462\|2020 | .................... | .................... | ..................... | .................... | .................... | .................... |
| F | 3223 | hCoV-19/USA/ME-HETL-J0052/2020\|EPI_ISL_513463\|2020 | .................... | .................... | ..................... | .................... | .................... | .................... |
| F | 3224 | hCoV-19/USA/ME-HETL-J0053/2020\|EPI_ISL_513464\|2020 | .................... | .................... | ..................... | .................... | .................... | .................... |
| F | 3225 | hCoV-19/USA/ME-HETL-J0054/2020\|EPI_ISL_513465\|2020 | .................... | .................... | ..................... | .................... | .................... | .................... |
| F | 3226 | hCoV-19/USA/ME-HETL-J0055/2020\|EPI_ISL_513466\|2020 | .................... | .................... | ..................... | .................... | .................... | .................... |
| F | 3227 | hCoV-19/USA/ME-HETL-J0056/2020\|EPI_ISL_513467\|2020 | .................... | .................... | ..................... | .................... | .................... | .................... |
| F | 3228 | hCoV-19/USA/ME-HETL-J0057/2020\|EPI_ISL_513468\|2020 | .................... | .................... | ..................... | .................... | .................... | .................... |
| F | 3229 | hCoV-19/USA/ME-HETL-J0058/2020\|EPI_ISL_513469\|2020 | .................... | .................... | ..................... | .................... | .................... | .................... |
| F | 3230 | hCoV-19/USA/ME-HETL-J0059/2020\|EPI_ISL_513470\|2020 | .................... | .................... | ..................... | .................... | .................... | .................... |
| F | 3231 | hCoV-19/USA/ME-HETL-J0060/2020\|EPI_ISL_513471\|2020 | .................... | .................... | ..................... | .................... | .................... | .................... |
| F | 3232 | hCoV-19/USA/ME-HETL-J0061/2020\|EPI_ISL_513472\|2020 | .................... | .................... | ..................... | .................... | .................... | .................... |
| F | 3233 | hCoV-19/USA/ME-HETL-J0062/2020\|EPI_ISL_513473\|2020 | .................... | .................... | ..................... | .................... | .................... | .................... |
| F | 3234 | hCoV-19/USA/ME-HETL-J0063/2020\|EPI_ISL_513474\|2020 | .................... | .................... | ..................... | .................... | .................... | .................... |
| F | 3235 | hCoV-19/USA/ME-HETL-J0064/2020\|EPI_ISL_513475\|2020 | .................... | .................... | ..................... | .................... | .................... | .................... |
| F | 3236 | hCoV-19/USA/ME-HETL-J0065/2020\|EPI_ISL_513476\|2020 | .................... | .................... | ..................... | .................... | .................... | .................... |
| F | 3238 | hCoV-19/USA/ME-HETL-J0067/2020\|EPI_ISL_513478\|2020 | .................... | .................... | ..................... | .................... | .................... | .................... |
| F | 3239 | hCoV-19/USA/ME-HETL-J0068/2020\|EPI_ISL_513479\|2020 | .................... | .................... | ..................... | .................... | .................... | .................... |
| F | 3240 | hCoV-19/USA/UN-CDC-7404/2020\|EPI_ISL_514062\|2020-0 | .................... | .................... | ..................... | .................... | .................... | .................... |
| F | 3241 | hCoV-19/USA/UN-CDC-7413/2020\|EPI_ISL_514063\|2020-0 | .................... | .................... | ..................... | .................... | .................... | .................... |
| F | 3242 | hCoV-19/USA/UN-CDC-7414/2020\|EPI_ISL_514064\|2020-0 | .................... | .................... | ..................... | .................... | .................... | .................... |
| F | 3243 | hCoV-19/USA/UN-CDC-7419/2020\|EPI_ISL_514065\|2020-0 | .................... | .................... | ..................... | .................... | .................... | .................... |
| F | 3244 | hCoV-19/USA/UN-CDC-7425/2020\|EPI_ISL_514066\|2020-0 | .................... | .................... | ..................... | .................... | .................... | .................... |
| F | 3245 | hCoV-19/USA/UN-CDC-7430/2020\|EPI_ISL_514067\|2020-0 | .................... | .................... | ..................... | .................... | .................... | .................... |
| F | 3246 | hCoV-19/USA/UN-CDC-7432/2020\|EPI_ISL_514068\|2020-0 | ............Y....... | .................... | ..................... | .................... | .................... | .................... |
| F | 3247 | hCoV-19/USA/UN-CDC-7433/2020\|EPI_ISL_514069\|2020-0 | .................... | .................... | ..................... | .................... | .................... | .................... |
| F | 3248 | hCoV-19/USA/UN-CDC-7441/2020\|EPI_ISL_514070\|2020-0 | .................... | .................... | ..................... | .................... | .................... | .................... |
| F | 3249 | hCoV-19/USA/UN-CDC-7462/2020\|EPI_ISL_514071\|2020-0 | .................... | .................... | ..................... | .................... | .................... | .................... |
| F | 3250 | hCoV-19/USA/UN-CDC-7467/2020\|EPI_ISL_514072\|2020-0 | .................... | .................... | ..................... | .................... | .................... | .................... |

F 3251 hCoV-19/USA/UN-CDC-7470/2020|EPI_ISL_514073|2020-0 .................... .................... .................... .................... .................... ....................
F 3252 hCoV-19/USA/UN-CDC-7480/2020|EPI_ISL_514074|2020-0 .................... .................... .................... .................... .................... ....................
F 3253 hCoV-19/USA/UN-CDC-7481/2020|EPI_ISL_514075|2020-0 .................... .................... .................... .................... .................... ....................
F 3254 hCoV-19/USA/UN-CDC-7493/2020|EPI_ISL_514077|2020-0 .................... .................... .................... .................... .................... ....................
F 3255 hCoV-19/USA/UN-CDC-7498/2020|EPI_ISL_514078|2020-0 .................... .................... .................... .................... .................... ....................
F 3256 hCoV-19/USA/UN-CDC-7520/2020|EPI_ISL_514079|2020-0 .................... .................... .................... .................... .................... ....................
F 3257 hCoV-19/USA/UN-CDC-7528/2020|EPI_ISL_514080|2020-0 .................... .................... .................... .................... .................... ....................
F 3258 hCoV-19/USA/UN-CDC-7529/2020|EPI_ISL_514081|2020-0 .................... .................... .................... .................... .................... ....................
F 3259 hCoV-19/USA/UN-CDC-7569/2020|EPI_ISL_514082|2020-0 .................... .................... .................... .................... .................... ....................
F 3260 hCoV-19/USA/UN-CDC-7618/2020|EPI_ISL_514083|2020-0 .................... .................... .................... .................... .................... ....................
F 3261 hCoV-19/USA/UN-CDC-7619/2020|EPI_ISL_514084|2020-0 .................... .................... .................... .................... .................... ....................
F 3262 hCoV-19/USA/UN-CDC-7625/2020|EPI_ISL_514085|2020-0 .................... .................... .................... .................... .................... ....................
F 3263 hCoV-19/USA/UN-CDC-7636/2020|EPI_ISL_514086|2020-0 .................... .................... .................... .................... .................... ....................
F 3264 hCoV-19/USA/UN-CDC-7645/2020|EPI_ISL_514087|2020-0 .................... .................... .................... .................... .................... ....................
F 3265 hCoV-19/USA/UN-CDC-7651/2020|EPI_ISL_514088|2020-0 .................... .................... .................... .................... .................... ....................
F 3266 hCoV-19/USA/UN-CDC-7652/2020|EPI_ISL_514089|2020-0 .................... .................... .................... .................... .................... ....................
F 3267 hCoV-19/USA/UN-CDC-7653/2020|EPI_ISL_514090|2020-0 .................... .................... .................... .................... .................... ....................
F 3268 hCoV-19/USA/UN-CDC-7657/2020|EPI_ISL_514091|2020-0 .................... .................... .................... .................... .................... ....................
F 3269 hCoV-19/USA/UN-CDC-7667/2020|EPI_ISL_514092|2020-0 .................... .................... .................... .................... .................... ....................
F 3270 hCoV-19/South Korea/KCDC2355/2020|EPI_ISL_514758|2 .................... .................... .................... .................... .................... ....................
F 3271 hCoV-19/South Korea/KCDC2356/2020|EPI_ISL_514759|2 .................... .................... .................... .................... .................... ....................
F 3272 hCoV-19/South Korea/KCDC2357/2020|EPI_ISL_514760|2 .................... .................... .................... .................... .................... ....................
F 3273 hCoV-19/South Korea/KCDC2358/2020|EPI_ISL_514761|2 .................... .................... .................... .................... .................... ....................
F 3274 hCoV-19/South Korea/KCDC2359/2020|EPI_ISL_514762|2 .................... .................... .................... .................... .................... ....................
F 3275 hCoV-19/South Korea/KCDC2360/2020|EPI_ISL_514763|2 .................... .................... .................... .................... .................... ....................
F 3276 hCoV-19/South Korea/KCDC2361/2020|EPI_ISL_514764|2 .................... .................... .................... .................... .................... ....................
F 3277 hCoV-19/South Korea/KCDC2362/2020|EPI_ISL_514765|2 .................... .................... .................... .................... .................... ....................
F 3278 hCoV-19/South Korea/KCDC2363/2020|EPI_ISL_514766|2 .................... .................... .................... .................... .................... ....................
F 3279 hCoV-19/South Korea/KCDC2364/2020|EPI_ISL_514767|2 .................... .................... .................... .................... .................... ....................
F 3280 hCoV-19/South Korea/KCDC2365/2020|EPI_ISL_514768|2 .................... .................... .................... .................... .................... ....................
F 3281 hCoV-19/South Korea/KCDC2366/2020|EPI_ISL_514769|2 .................... .................... .................... .................... .................... ....................
F 3282 hCoV-19/South Korea/KCDC2367/2020|EPI_ISL_514770|2 .................... .................... .................... .................... .................... ....................
F 3283 hCoV-19/South Korea/KCDC2368/2020|EPI_ISL_514771|2 .................... .................... .................... .................... .................... ....................
F 3284 hCoV-19/South Korea/KCDC2369/2020|EPI_ISL_514772|2 .................... .................... .................... .................... .................... ....................
F 3285 hCoV-19/South Korea/KCDC2370/2020|EPI_ISL_514773|2 .................... .................... .................... .................... .................... ....................
F 3286 hCoV-19/South Korea/KCDC2371/2020|EPI_ISL_514774|2 .................... .................... .................... .................... .................... ....................
F 3287 hCoV-19/South Korea/KCDC2372/2020|EPI_ISL_514775|2 .................... .................... .................... .................... .................... ....................
F 3288 hCoV-19/South Korea/KCDC2373/2020|EPI_ISL_514776|2 .................... .................... .................... .................... .................... ....................
F 3289 hCoV-19/South Korea/KCDC2374/2020|EPI_ISL_514777|2 .................... .................... .................... .................... .................... ....................
F 3290 hCoV-19/South Korea/KCDC2375/2020|EPI_ISL_514778|2 .................... .................... .................... .................... .................... ....................
F 3291 hCoV-19/South Korea/KCDC2376/2020|EPI_ISL_514779|2 .................... .................... .................... .................... .................... ....................
F 3292 hCoV-19/South Korea/KCDC2377/2020|EPI_ISL_514780|2 .................... .................... .................... .................... .................... ....................
F 3293 hCoV-19/South Korea/KCDC2378/2020|EPI_ISL_514781|2 .................... .................... .................... .................... .................... ....................
F 3294 hCoV-19/South Korea/KCDC2379/2020|EPI_ISL_514782|2 .................... .................... .................... .................... .................... ....................
F 3295 hCoV-19/South Korea/KCDC2380/2020|EPI_ISL_514783|2 .................... .................... .................... .................... .................... ....................
F 3296 hCoV-19/South Korea/KCDC2381/2020|EPI_ISL_514784|2 .................... .................... .................... .................... .................... ....................
F 3297 hCoV-19/South Korea/KCDC2382/2020|EPI_ISL_514785|2 .................... .................... .................... .................... .................... ....................
F 3298 hCoV-19/South Korea/KCDC2383/2020|EPI_ISL_514786|2 .................... .................... .................... .................... .................... ....................
F 3299 hCoV-19/South Korea/KCDC2384/2020|EPI_ISL_514787|2 .................... .................... .................... .................... .................... ....................
F 3300 hCoV-19/South Korea/KCDC2385/2020|EPI_ISL_514788|2 .................... .................... .................... .................... .................... ....................
F 3301 hCoV-19/South Korea/KCDC2386/2020|EPI_ISL_514789|2 .................... .................... .................... .................... .................... ....................
F 3302 hCoV-19/South Korea/KCDC2387/2020|EPI_ISL_514790|2 .................... .................... .................... .................... .................... ....................
F 3303 hCoV-19/South Korea/KCDC2388/2020|EPI_ISL_514791|2 .................... .................... .................... .................... .................... ....................
F 3304 hCoV-19/South Korea/KCDC2389/2020|EPI_ISL_514792|2 .................... .................... .................... .................... .................... ....................
F 3305 hCoV-19/South Korea/KCDC2390/2020|EPI_ISL_514793|2 .................... .................... .................... .................... .................... ....................
F 3306 hCoV-19/South Korea/KCDC2391/2020|EPI_ISL_514794|2 .................... .................... .................... .................... .................... ....................
F 3307 hCoV-19/South Korea/KCDC2392/2020|EPI_ISL_514795|2 .................... .................... .................... .................... .................... ....................
F 3308 hCoV-19/South Korea/KCDC2393/2020|EPI_ISL_514796|2 .................... .................... .................... .................... .................... ....................
F 3309 hCoV-19/South Korea/KCDC2394/2020|EPI_ISL_514797|2 .................... .................... .................... .................... .................... ....................
F 3310 hCoV-19/South Korea/KCDC2395/2020|EPI_ISL_514798|2 .................... .................... .................... .................... .................... ....................
F 3311 hCoV-19/South Korea/KCDC2396/2020|EPI_ISL_514799|2 .................... .................... .................... .................... .................... ....................
F 3312 hCoV-19/USA/NV-NSPHL-A0108/2020|EPI_ISL_515381|202 .................... .................... .................... .................... .................... ....................
F 3313 hCoV-19/USA/NV-NSPHL-A0111/2020|EPI_ISL_515382|202 .................... .................... .................... .................... .................... ....................
F 3314 hCoV-19/USA/NV-NSPHL-A0113/2020|EPI_ISL_515384|202 .................... .................... .................... .................... .................... ....................
F 3315 hCoV-19/USA/NV-NSPHL-A0114/2020|EPI_ISL_515385|202 .................... .................... .................... .................... .................... ....................
F 3316 hCoV-19/USA/NV-NSPHL-A0115/2020|EPI_ISL_515386|202 .................... .................... .................... .................... .................... ....................
F 3317 hCoV-19/USA/NV-NSPHL-A0116/2020|EPI_ISL_515387|202 .................... .................... .................... .................... .................... ....................
F 3318 hCoV-19/USA/NV-NSPHL-A0117/2020|EPI_ISL_515388|202 .................... .................... .................... .................... .................... ....................
F 3319 hCoV-19/USA/NV-NSPHL-A0118/2020|EPI_ISL_515389|202 .................... .................... .................... .................... .................... ....................
F 3320 hCoV-19/USA/NV-NSPHL-A0119/2020|EPI_ISL_515390|202 .................... .................... .................... .................... .................... ....................
F 3321 hCoV-19/USA/NV-NSPHL-A0120/2020|EPI_ISL_515391|202 .................... .................... .................... .................... .................... ....................
F 3322 hCoV-19/USA/NV-NSPHL-A0121/2020|EPI_ISL_515392|202 .................... .................... .................... .................... .................... ....................
F 3323 hCoV-19/USA/NV-NSPHL-A0122/2020|EPI_ISL_515393|202 .................... .................... .................... .................... .................... ....................
F 3324 hCoV-19/USA/NV-NSPHL-A0123/2020|EPI_ISL_515394|202 .................... .................... .................... .................... .................... ....................
F 3325 hCoV-19/env/USA/NV-NSPHL-A0128/2020|EPI_ISL_515396 .................... .................... .................... .................... .................... ....................
F 3326 hCoV-19/USA/NV-NSPHL-A0129/2020|EPI_ISL_515397|202 .................... .................... .................... .................... .................... ....................
F 3327 hCoV-19/env/USA/NV-NSPHL-A0130/2020|EPI_ISL_515398 .................... .................... .................... .................... .................... ....................
F 3328 hCoV-19/USA/NV-NSPHL-A0131/2020|EPI_ISL_515399|202 .................... .................... .................... .................... .................... ....................
F 3329 hCoV-19/USA/WA-S1874/2020|EPI_ISL_496300|2020-05-2 .................... .................... .................... .................... .................... ....................
F 3330 hCoV-19/USA/WA-S1875/2020|EPI_ISL_496301|2020-05-2 .................... .................... .................... .................... .................... ....................

| | | | | | | | | |
|---|---|---|---|---|---|---|---|---|
| F | 3331 | hCoV-19/USA/WA-S1876/2020\|EPI_ISL_496302\|2020-05-2 | ···················· | ···················· | ····················· | ···················· | ···················· | ···················· |
| F | 3332 | hCoV-19/USA/WA-S1877/2020\|EPI_ISL_496303\|2020-05-2 | ···················· | ···················· | ····················· | ···················· | ···················· | ···················· |
| F | 3333 | hCoV-19/USA/WA-S1878/2020\|EPI_ISL_496304\|2020-05-2 | ···················· | ···················· | ····················· | ···················· | ···················· | ···················· |
| F | 3334 | hCoV-19/USA/WA-S1879/2020\|EPI_ISL_496305\|2020-05-2 | ···················· | ···················· | ····················· | ···················· | ···················· | ···················· |
| F | 3335 | hCoV-19/USA/WA-S1880/2020\|EPI_ISL_496306\|2020-05-2 | ···················· | ···················· | ····················· | ···················· | ···················· | ···················· |
| F | 3336 | hCoV-19/USA/WA-S1881/2020\|EPI_ISL_496307\|2020-05-2 | ···················· | ···················· | ····················· | ···················· | ···················· | ···················· |
| F | 3337 | hCoV-19/USA/WA-S1882/2020\|EPI_ISL_496308\|2020-05-2 | ···················· | ···················· | ····················· | ···················· | ···················· | ···················· |
| F | 3338 | hCoV-19/USA/WA-S1883/2020\|EPI_ISL_496309\|2020-05-2 | ···················· | ···················· | ····················· | ···················· | ···················· | ···················· |
| F | 3339 | hCoV-19/USA/WA-S1884/2020\|EPI_ISL_496310\|2020-05-2 | ···················· | ···················· | ····················· | ···················· | ···················· | ···················· |
| F | 3340 | hCoV-19/USA/WA-S1889/2020\|EPI_ISL_496315\|2020-05-2 | ···················· | ···················· | ····················· | ···················· | ···················· | ···················· |
| F | 3341 | hCoV-19/USA/WA-S1890/2020\|EPI_ISL_496316\|2020-05-2 | ···················· | ···················· | ····················· | ···················· | ···················· | ···················· |
| F | 3342 | hCoV-19/USA/WA-S1891/2020\|EPI_ISL_496317\|2020-05-2 | ···················· | ···················· | ····················· | ···················· | ···················· | ···················· |
| F | 3343 | hCoV-19/USA/WA-S1892/2020\|EPI_ISL_496318\|2020-05-2 | ···················· | ···················· | ····················· | ···················· | ···················· | ···················· |
| F | 3344 | hCoV-19/USA/WA-S1893/2020\|EPI_ISL_496319\|2020-05-2 | ···················· | ···················· | ····················· | ···················· | ···················· | ···················· |
| F | 3345 | hCoV-19/USA/WA-S1894/2020\|EPI_ISL_496320\|2020-05-2 | ···················· | ···················· | ····················· | ···················· | ···················· | ···················· |
| F | 3346 | hCoV-19/USA/WA-S1895/2020\|EPI_ISL_496321\|2020-05-2 | ···················· | ···················· | ····················· | ···················· | ···················· | ···················· |
| F | 3347 | hCoV-19/USA/WA-S1907/2020\|EPI_ISL_496333\|2020-05-2 | ···················· | ···················· | ····················· | ···················· | ···················· | ···················· |
| F | 3348 | hCoV-19/USA/WA-S1908/2020\|EPI_ISL_496334\|2020-05-2 | ···················· | ···················· | ····················· | ···················· | ···················· | ···················· |
| F | 3349 | hCoV-19/USA/WA-S1909/2020\|EPI_ISL_496335\|2020-05-2 | ···················· | ···················· | ····················· | ···················· | ···················· | ···················· |
| F | 3350 | hCoV-19/USA/WA-S1910/2020\|EPI_ISL_496336\|2020-05-2 | ···················· | ···················· | ····················· | ···················· | ···················· | ···················· |
| F | 3351 | hCoV-19/USA/WA-S1911/2020\|EPI_ISL_496337\|2020-05-2 | ···················· | ···················· | ····················· | ···················· | ···················· | ···················· |
| F | 3352 | hCoV-19/USA/WA-S1912/2020\|EPI_ISL_496338\|2020-05-2 | ···················· | ···················· | ····················· | ···················· | ···················· | ···················· |
| F | 3353 | hCoV-19/Mexico/BCN-ALSR-1486/2020\|EPI_ISL_496350\|2 | ···················· | ···················· | ····················· | ···················· | ···················· | ···················· |
| F | 3354 | hCoV-19/USA/WA-S2172/2020\|EPI_ISL_497600\|2020-06-0 | ···················· | ···················· | ····················· | ···················· | ···················· | ···················· |
| F | 3355 | hCoV-19/USA/WA-S2173/2020\|EPI_ISL_497601\|2020-06-0 | ···················· | ···················· | ····················· | ···················· | ···················· | ···················· |
| F | 3356 | hCoV-19/USA/WA-S2175/2020\|EPI_ISL_497603\|2020-06-0 | ···················· | ···················· | ····················· | ···················· | ···················· | ···················· |
| F | 3357 | hCoV-19/USA/WA-S2176/2020\|EPI_ISL_497604\|2020-06-0 | ···················· | ···················· | ····················· | ···················· | ···················· | ···················· |
| F | 3358 | hCoV-19/USA/WA-S2177/2020\|EPI_ISL_497605\|2020-06-0 | ···················· | ···················· | ····················· | ···················· | ···················· | ···················· |
| F | 3359 | hCoV-19/USA/WA-S2179/2020\|EPI_ISL_497607\|2020-06-0 | ···················· | ···················· | ····················· | ···················· | ···················· | ···················· |
| F | 3360 | hCoV-19/USA/WA-S2180/2020\|EPI_ISL_497608\|2020-06-0 | ···················· | ···················· | ····················· | ···················· | ···················· | ···················· |
| F | 3361 | hCoV-19/USA/WA-S2181/2020\|EPI_ISL_497609\|2020-06-0 | ···················· | ···················· | ····················· | ···················· | ···················· | ···················· |
| F | 3362 | hCoV-19/USA/WA-S2182/2020\|EPI_ISL_497610\|2020-06-0 | ···················· | ···················· | ····················· | ···················· | ···················· | ···················· |
| F | 3363 | hCoV-19/USA/WA-S2183/2020\|EPI_ISL_497611\|2020-06-0 | ···················· | ···················· | ····················· | ···················· | ···················· | ···················· |
| F | 3364 | hCoV-19/USA/WA-S2184/2020\|EPI_ISL_497612\|2020-06-0 | ···················· | ···················· | ····················· | ···················· | ···················· | ···················· |
| F | 3365 | hCoV-19/USA/WA-S2185/2020\|EPI_ISL_497613\|2020-06-0 | ···················· | ···················· | ····················· | ···················· | ···················· | ···················· |
| F | 3366 | hCoV-19/USA/WA-S2186/2020\|EPI_ISL_497614\|2020-06-0 | ···················· | ···················· | ····················· | ···················· | ···················· | ···················· |
| F | 3367 | hCoV-19/USA/WA-S2187/2020\|EPI_ISL_497615\|2020-06-0 | ···················· | ···················· | ····················· | ···················· | ···················· | ···················· |
| F | 3368 | hCoV-19/USA/WA-S2188/2020\|EPI_ISL_497616\|2020-06-0 | ···················· | ···················· | ····················· | ···················· | ···················· | ···················· |
| F | 3369 | hCoV-19/USA/WA-S2189/2020\|EPI_ISL_497617\|2020-06-0 | ···················· | ···················· | ····················· | ···················· | ···················· | ···················· |
| F | 3370 | hCoV-19/USA/WA-S2190/2020\|EPI_ISL_497618\|2020-06-0 | ···················· | ···················· | ····················· | ···················· | ···················· | ···················· |
| F | 3371 | hCoV-19/USA/WA-S2191/2020\|EPI_ISL_497619\|2020-06-0 | ···················· | ···················· | ····················· | ···················· | ···················· | ···················· |
| F | 3372 | hCoV-19/USA/WA-S2192/2020\|EPI_ISL_497620\|2020-06-0 | ···················· | ···················· | ····················· | ···················· | ···················· | ···················· |
| F | 3373 | hCoV-19/USA/WA-S2195/2020\|EPI_ISL_497623\|2020-06-0 | ···················· | ···················· | ····················· | ···················· | ···················· | ···················· |
| F | 3374 | hCoV-19/USA/WA-S2196/2020\|EPI_ISL_497624\|2020-06-0 | ···················· | ···················· | ····················· | ···················· | ···················· | ···················· |
| F | 3375 | hCoV-19/USA/WA-S2198/2020\|EPI_ISL_497626\|2020-06-0 | ···················· | ···················· | ····················· | ···················· | ···················· | ···················· |
| F | 3376 | hCoV-19/USA/WA-S2200/2020\|EPI_ISL_497628\|2020-06-0 | ···················· | ···················· | ····················· | ···················· | ···················· | ···················· |
| F | 3377 | hCoV-19/USA/WA-S2201/2020\|EPI_ISL_497629\|2020-06-0 | ···················· | ···················· | ····················· | ···················· | ···················· | ···················· |
| F | 3378 | hCoV-19/USA/WA-S2202/2020\|EPI_ISL_497630\|2020-06-0 | ···················· | ···················· | ····················· | ···················· | ···················· | ···················· |
| F | 3379 | hCoV-19/USA/WA-S2203/2020\|EPI_ISL_497631\|2020-06-0 | ···················· | ···················· | ····················· | ···················· | ···················· | ···················· |
| F | 3380 | hCoV-19/USA/WA-S2204/2020\|EPI_ISL_497632\|2020-06-0 | ···················· | ···················· | ····················· | ···················· | ···················· | ···················· |
| F | 3381 | hCoV-19/USA/WA-S2205/2020\|EPI_ISL_497633\|2020-06-0 | ···················· | ···················· | ····················· | ···················· | ···················· | ···················· |
| F | 3382 | hCoV-19/USA/WA-S2206/2020\|EPI_ISL_497634\|2020-06-0 | ···················· | ···················· | ····················· | ···················· | ···················· | ···················· |
| F | 3383 | hCoV-19/USA/WA-S2208/2020\|EPI_ISL_497636\|2020-06-0 | ···················· | ···················· | ····················· | ···················· | ···················· | ···················· |
| F | 3384 | hCoV-19/USA/WA-S2209/2020\|EPI_ISL_497637\|2020-06-0 | ···················· | ···A················ | ····················· | ···················· | ···················· | ···················· |
| F | 3385 | hCoV-19/USA/WA-S2210/2020\|EPI_ISL_497638\|2020-06-0 | ···················· | ···················· | ····················· | ···················· | ···················· | ···················· |
| F | 3386 | hCoV-19/USA/WA-S2211/2020\|EPI_ISL_497639\|2020-06-0 | ···················· | ···················· | ····················· | ···················· | ···················· | ···················· |
| F | 3387 | hCoV-19/USA/WA-S2212/2020\|EPI_ISL_497640\|2020-06-0 | ···················· | ···················· | ····················· | ···················· | ···················· | ···················· |
| F | 3388 | hCoV-19/USA/WA-S2213/2020\|EPI_ISL_497641\|2020-06-0 | ···················· | ···················· | ····················· | ···················· | ···················· | ···················· |
| F | 3389 | hCoV-19/USA/WA-S2218/2020\|EPI_ISL_497646\|2020-06-0 | ···················· | ···················· | ····················· | ···················· | ···················· | ···················· |
| F | 3390 | hCoV-19/USA/WA-S2219/2020\|EPI_ISL_497647\|2020-06-0 | ···················· | ···················· | ····················· | ···················· | ···················· | ···················· |
| F | 3391 | hCoV-19/USA/WA-S2220/2020\|EPI_ISL_497648\|2020-06-0 | ···················· | ···················· | ····················· | ···················· | ···················· | ···················· |
| F | 3392 | hCoV-19/USA/WA-S2222/2020\|EPI_ISL_497650\|2020-06-0 | ···················· | ···················· | ····················· | ···················· | ···················· | ···················· |
| F | 3393 | hCoV-19/USA/WA-S2223/2020\|EPI_ISL_497651\|2020-06-0 | ···················· | ···················· | ····················· | ···················· | ···················· | ···················· |
| F | 3394 | hCoV-19/USA/WA-S2224/2020\|EPI_ISL_497652\|2020-06-0 | ···················· | ···················· | ····················· | ···················· | ···················· | ···················· |
| F | 3395 | hCoV-19/USA/WA-S2236/2020\|EPI_ISL_497664\|2020-06-0 | ···················· | ···················· | ····················· | ···················· | ···················· | ···················· |
| F | 3396 | hCoV-19/USA/WA-S2237/2020\|EPI_ISL_497665\|2020-06-0 | ···················· | ···················· | ····················· | ···················· | ···················· | ···················· |
| F | 3397 | hCoV-19/USA/WA-S2239/2020\|EPI_ISL_497667\|2020-06-0 | ···················· | ···················· | ····················· | ···················· | ···················· | ···················· |
| F | 3398 | hCoV-19/USA/WA-S2240/2020\|EPI_ISL_497668\|2020-06-0 | ···················· | ···················· | ····················· | ···················· | ···················· | ···················· |
| F | 3399 | hCoV-19/USA/WA-S2241/2020\|EPI_ISL_497669\|2020-06-0 | ···················· | ···················· | ····················· | ···················· | ···················· | ···················· |
| F | 3400 | hCoV-19/USA/WA-S2242/2020\|EPI_ISL_497670\|2020-06-0 | ···················· | ···················· | ····················· | ···················· | ···················· | ···················· |
| F | 3401 | hCoV-19/USA/WA-S2243/2020\|EPI_ISL_497671\|2020-06-0 | ···················· | ···················· | ····················· | ···················· | ···················· | ···················· |
| F | 3402 | hCoV-19/USA/WA-S2244/2020\|EPI_ISL_497672\|2020-06-0 | ···················· | ···················· | ····················· | ···················· | ···················· | ···················· |
| F | 3403 | hCoV-19/USA/WA-S2245/2020\|EPI_ISL_497673\|2020-06-0 | ···················· | ···················· | ····················· | ···················· | ···················· | ···················· |
| F | 3404 | hCoV-19/USA/WA-S2246/2020\|EPI_ISL_497674\|2020-06-0 | ···················· | ···A················ | ····················· | ···················· | ···················· | ···················· |
| F | 3405 | hCoV-19/USA/WA-S2247/2020\|EPI_ISL_497675\|2020-06-0 | ···················· | ···A················ | ····················· | ···················· | ···················· | ···················· |
| F | 3406 | hCoV-19/USA/WA-S2248/2020\|EPI_ISL_497676\|2020-06-0 | ···················· | ···················· | ····················· | ···················· | ···················· | ···················· |
| F | 3407 | hCoV-19/USA/WA-S2249/2020\|EPI_ISL_497677\|2020-06-0 | ···················· | ···················· | ····················· | ···················· | ···················· | ···················· |
| F | 3408 | hCoV-19/USA/WA-S2251/2020\|EPI_ISL_497679\|2020-06-0 | ···················· | ···················· | ····················· | ···················· | ···················· | ···················· |
| F | 3409 | hCoV-19/USA/WA-S2252/2020\|EPI_ISL_497680\|2020-06-0 | ···················· | ···················· | ····················· | ···················· | ···················· | ···················· |
| F | 3410 | hCoV-19/USA/WA-S2253/2020\|EPI_ISL_497681\|2020-06-0 | ···················· | ···················· | ····················· | ···················· | ···················· | ···················· |

| | | | | | | | | | |
|---|---|---|---|---|---|---|---|---|---|
| F | 3411 | hCoV-19/USA/WA-S2254/2020\|EPI_ISL_497682\|2020-06-0 | ···················· | ····A··············· | ···················· | ···················· | ···················· | ···················· | |
| F | 3412 | hCoV-19/USA/WA-S2255/2020\|EPI_ISL_497683\|2020-06-0 | ···················· | ···················· | ···················· | ···················· | ···················· | ···················· | |
| F | 3413 | hCoV-19/USA/WA-S2256/2020\|EPI_ISL_497684\|2020-06-0 | ···················· | ···················· | ···················· | ···················· | ···················· | ···················· | |
| F | 3414 | hCoV-19/USA/WA-S2257/2020\|EPI_ISL_497685\|2020-06-0 | ···················· | ···················· | ···················· | ···················· | ···················· | ···················· | |
| F | 3415 | hCoV-19/Bangladesh/BCSIR-NILMRC-319/2020\|EPI_ISL_4 | ···················· | ···················· | ···················· | ···················· | ···················· | ···················· | |
| F | 3416 | hCoV-19/Bangladesh/BCSIR-NILMRC-321/2020\|EPI_ISL_4 | ···················· | ···················· | ···················· | ···················· | ···················· | ···················· | |
| F | 3417 | hCoV-19/England/LIVE-A4FC5/2020\|EPI_ISL_500100\|202 | ···················· | ···················· | ···················· | ···················· | ···················· | ···················· | |
| F | 3418 | hCoV-19/England/LIVE-A5058/2020\|EPI_ISL_500101\|202 | ···················· | ···················· | ···················· | ···················· | ···················· | ···················· | |
| F | 3419 | hCoV-19/England/LIVE-A5067/2020\|EPI_ISL_500102\|202 | ···················· | ···················· | ···················· | ···················· | ···················· | ···················· | |
| F | 3420 | hCoV-19/England/LIVE-A5076/2020\|EPI_ISL_500103\|202 | ···················· | ···················· | ···················· | ···················· | ···················· | ···················· | |
| F | 3422 | hCoV-19/England/LIVE-A50C1/2020\|EPI_ISL_500105\|202 | ···················· | ···················· | ···················· | ···················· | ···················· | ···················· | |
| F | 3425 | hCoV-19/England/LIVE-A5498/2020\|EPI_ISL_500119\|202 | ···················· | ···················· | ···················· | ···················· | ···················· | ···················· | |
| F | 3427 | hCoV-19/England/LIVE-A54C5/2020\|EPI_ISL_500121\|202 | ···················· | ···················· | ···················· | ···················· | ···················· | ···················· | |
| F | 3428 | hCoV-19/England/LIVE-A54D4/2020\|EPI_ISL_500122\|202 | ···················· | ···················· | ···················· | ···················· | ···················· | ···················· | |
| F | 3431 | hCoV-19/England/LIVE-A5559/2020\|EPI_ISL_500125\|202 | ···················· | ···················· | ···················· | ···················· | ···················· | ···················· | |
| F | 3433 | hCoV-19/England/LIVE-A5586/2020\|EPI_ISL_500127\|202 | ···················· | ···················· | ···················· | ···················· | ···················N | ···················· | 1 |
| F | 3434 | hCoV-19/Brazil/PE-AMU0036/2020\|EPI_ISL_502779\|2020 | ···················· | ···················· | ···················· | ···················· | ···················· | ···················· | |
| F | 3435 | hCoV-19/USA/MI-MDHHS-SC21348/2020\|EPI_ISL_507602\|2 | ···················· | ···················· | ···················· | ···················· | ···················· | ···················· | |
| F | 3436 | hCoV-19/USA/MI-MDHHS-SC21349/2020\|EPI_ISL_507603\|2 | ···················· | ···················· | ···················· | ···················· | ···················· | ···················· | |
| F | 3437 | hCoV-19/USA/MI-MDHHS-SC21350/2020\|EPI_ISL_507604\|2 | ···················· | ···················· | ···················· | ···················· | ···················· | ···················· | |
| F | 3438 | hCoV-19/USA/MI-MDHHS-SC21351/2020\|EPI_ISL_507605\|2 | ···················· | ···················· | ···················· | ···················· | ···················· | ···················· | |
| F | 3439 | hCoV-19/USA/MI-MDHHS-SC21352/2020\|EPI_ISL_507606\|2 | ···················· | ···················· | ···················· | ···················· | ···················· | ···················· | |
| F | 3440 | hCoV-19/USA/MI-MDHHS-SC21353/2020\|EPI_ISL_507607\|2 | ···················· | ···················· | ···················· | ···················· | ···················· | ···················· | |
| F | 3441 | hCoV-19/USA/MI-MDHHS-SC21354/2020\|EPI_ISL_507608\|2 | ···················· | ···················· | ···················· | ···················· | ···················· | ···················· | |
| F | 3442 | hCoV-19/USA/MI-MDHHS-SC21355/2020\|EPI_ISL_507609\|2 | ···················· | ···················· | ···················· | ···················· | ···················· | ···················· | |
| F | 3443 | hCoV-19/USA/MI-MDHHS-SC21356/2020\|EPI_ISL_507610\|2 | ···················· | ···················· | ···················· | ···················· | ···················· | ···················· | |
| F | 3444 | hCoV-19/USA/MI-MDHHS-SC21357/2020\|EPI_ISL_507611\|2 | ···················· | ···················· | ···················· | ···················· | ···················· | ···················· | |
| F | 3445 | hCoV-19/USA/MI-MDHHS-SC21358/2020\|EPI_ISL_507612\|2 | ···················· | ···················· | ···················· | ···················· | ···················· | ···················· | |
| F | 3446 | hCoV-19/USA/MI-MDHHS-SC21359/2020\|EPI_ISL_507613\|2 | ···················· | ···················· | ···················· | ···················· | ···················· | ···················· | |
| F | 3447 | hCoV-19/USA/MI-MDHHS-SC21360/2020\|EPI_ISL_507614\|2 | ···················· | ···················· | ···················· | ···················· | ···················· | ···················· | |
| F | 3448 | hCoV-19/USA/MI-MDHHS-SC21361/2020\|EPI_ISL_507615\|2 | ···················· | ···················· | ···················· | ···················· | ···················· | ···················· | |
| F | 3449 | hCoV-19/USA/MI-MDHHS-SC21362/2020\|EPI_ISL_507616\|2 | ···················· | ···················· | ···················· | ···················· | ···················· | ···················· | |
| F | 3450 | hCoV-19/USA/MI-MDHHS-SC21363/2020\|EPI_ISL_507617\|2 | ···················· | ···················· | ···················· | ···················· | ···················· | ···················· | |
| F | 3451 | hCoV-19/USA/MI-MDHHS-SC21364/2020\|EPI_ISL_507618\|2 | ···················· | ···················· | ···················· | ···················· | ···················· | ···················· | |
| F | 3452 | hCoV-19/USA/MI-MDHHS-SC21365/2020\|EPI_ISL_507619\|2 | ···················· | ···················· | ···················· | ···················· | ···················· | ···················· | |
| F | 3453 | hCoV-19/USA/MI-MDHHS-SC21366/2020\|EPI_ISL_507620\|2 | ···················· | ···················· | ···················· | ···················· | ···················· | ···················· | |
| F | 3454 | hCoV-19/USA/MI-MDHHS-SC21367/2020\|EPI_ISL_507621\|2 | ···················· | ···················· | ···················· | ···················· | ···················· | ···················· | |
| F | 3455 | hCoV-19/USA/MI-MDHHS-SC21368/2020\|EPI_ISL_507622\|2 | ···················· | ···················· | ···················· | ···················· | ···················· | ···················· | |
| F | 3456 | hCoV-19/USA/MI-MDHHS-SC21369/2020\|EPI_ISL_507623\|2 | ···················· | ···················· | ···················· | ···················· | ···················· | ···················· | |
| F | 3457 | hCoV-19/France/ARA-16220/2020\|EPI_ISL_508906\|2020- | ···················· | ···················· | ···················· | ···················· | ···················· | ···················· | |
| F | 3458 | hCoV-19/France/ARA-16251/2020\|EPI_ISL_508907\|2020- | ···················· | ···················· | ···················· | ···················· | ···················· | ···················· | |
| F | 3459 | hCoV-19/France/ARA-16322/2020\|EPI_ISL_508908\|2020- | ···················· | ···················· | ···················· | ···················· | ···················· | ···················· | |
| F | 3460 | hCoV-19/France/OCC-MP01829/2020\|EPI_ISL_508909\|202 | ···················· | ···················· | ···················· | ···················· | ···················· | ···················· | |
| F | 3461 | hCoV-19/France/NAQ-NA01745/2020\|EPI_ISL_508910\|202 | ···················· | ···················· | ···················· | ···················· | ···················· | ···················· | |
| F | 3462 | hCoV-19/France/PAC-01926/2020\|EPI_ISL_508911\|2020- | ···················· | ···················· | ···················· | ···················· | ···················· | ···················· | |
| F | 3463 | hCoV-19/France/ARA-20756/2020\|EPI_ISL_508912\|2020- | ···················· | ···················· | ···················· | ···················· | ···················· | ···················· | |
| F | 3464 | hCoV-19/France/ARA-20768/2020\|EPI_ISL_508913\|2020- | ···················· | ···················· | ···················· | ···················· | ···················· | ···················· | |
| F | 3465 | hCoV-19/France/ARA-20829/2020\|EPI_ISL_508914\|2020- | ···················· | ···················· | ···················· | ···················· | ···················· | ···················· | |
| F | 3466 | hCoV-19/France/ARA-20976/2020\|EPI_ISL_508915\|2020- | ···················· | ···················· | ···················· | ···················· | ···················· | ···················· | |
| F | 3467 | hCoV-19/France/ARA-21170/2020\|EPI_ISL_508916\|2020- | ···················· | ···················· | ···················· | ···················· | ···················· | ···················· | |
| F | 3468 | hCoV-19/France/ARA-2221/2020\|EPI_ISL_508917\|2020-0 | ···················· | ···················· | ···················· | ···················· | ···················· | ···················· | |
| F | 3469 | hCoV-19/France/ARA-22592/2020\|EPI_ISL_508918\|2020- | ···················· | ···················· | ···················· | ···················· | ···················· | ···················· | |
| F | 3470 | hCoV-19/France/ARA-22670/2020\|EPI_ISL_508921\|2020- | ···················· | ···················· | ···················· | ···················· | ···················· | ···················· | |
| F | 3471 | hCoV-19/France/ARA-22864/2020\|EPI_ISL_508922\|2020- | ···················· | ···················· | ···················· | ···················· | ···················· | ···················· | |
| F | 3472 | hCoV-19/France/ARA-23001/2020\|EPI_ISL_508923\|2020- | ···················· | ···················· | ···················· | ···················· | ···················· | ···················· | |
| F | 3473 | hCoV-19/France/ARA-23076/2020\|EPI_ISL_508924\|2020- | ···················· | ···················· | ···················· | ···················· | ···················· | ···················· | |
| F | 3474 | hCoV-19/France/ARA-23092/2020\|EPI_ISL_508925\|2020- | ···················· | ···················· | ···················· | ···················· | ···················· | ···················· | |
| F | 3475 | hCoV-19/France/ARA-23121/2020\|EPI_ISL_508926\|2020- | ···················· | ···················· | ···················· | ···················· | ···················· | ···················· | |
| F | 3476 | hCoV-19/France/ARA-23544/2020\|EPI_ISL_508927\|2020- | ···················· | ···················· | ···················· | ···················· | ···················· | ···················· | |
| F | 3477 | hCoV-19/France/ARA-23813/2020\|EPI_ISL_508928\|2020- | ···················· | ···················· | ···················· | ···················· | ···················· | ···················· | |
| F | 3478 | hCoV-19/France/ARA-23893/2020\|EPI_ISL_508929\|2020- | ···················· | ···················· | ···················· | ···················· | ···················· | ···················· | |
| F | 3479 | hCoV-19/France/ARA-23943/2020\|EPI_ISL_508930\|2020- | ···················· | ···················· | ···················· | ···················· | ···················· | ···················· | |
| F | 3480 | hCoV-19/France/ARA-24023/2020\|EPI_ISL_508931\|2020- | ···················· | ···················· | ···················· | ···················· | ···················· | ···················· | |
| F | 3481 | hCoV-19/France/ARA-24115/2020\|EPI_ISL_508932\|2020- | ···················· | ···················· | ···················· | ···················· | ···················· | ···················· | |
| F | 3482 | hCoV-19/France/ARA-05288/2020\|EPI_ISL_508933\|2020- | ···················· | ···················· | ···················· | ···················· | ···················· | ···················· | |
| F | 3483 | hCoV-19/France/ARA-0591/2020\|EPI_ISL_508934\|2020-0 | ···················· | ···················· | ···················· | ···················· | ···················· | ···················· | |
| F | 3484 | hCoV-19/France/ARA-0592/2020\|EPI_ISL_508935\|2020-0 | ···················· | ···················· | ···················· | ···················· | ···················· | ···················· | |
| F | 3485 | hCoV-19/France/ARA-06032/2020\|EPI_ISL_508936\|2020- | ···················· | ···················· | ···················· | ···················· | ···················· | ···················· | |
| F | 3486 | hCoV-19/France/ARA-06923/2020\|EPI_ISL_508937\|2020- | ···················· | ···················· | ···················· | ···················· | ···················· | ···················· | |
| F | 3487 | hCoV-19/France/ARA-07055/2020\|EPI_ISL_508938\|2020- | ···················· | ···················· | ···················· | ···················· | ···················· | ···················· | |
| F | 3488 | hCoV-19/France/ARA-07148/2020\|EPI_ISL_508939\|2020- | ···················· | ···················· | ···················· | ···················· | ···················· | ···················· | |
| F | 3489 | hCoV-19/France/ARA-07169/2020\|EPI_ISL_508940\|2020- | ···················· | ···················· | ···················· | ···················· | ···················· | ···················· | |
| F | 3490 | hCoV-19/France/ARA-07314/2020\|EPI_ISL_508942\|2020- | ···················· | ···················· | ···················· | ···················· | ···················· | ···················· | |
| F | 3491 | hCoV-19/France/ARA-07722/2020\|EPI_ISL_508943\|2020- | ···················· | ···················· | ···················· | ···················· | ···················· | ···················· | |
| F | 3492 | hCoV-19/France/ARA-07731/2020\|EPI_ISL_508944\|2020- | ···················· | ···················· | ···················· | ···················· | ···················· | ···················· | |
| F | 3493 | hCoV-19/France/ARA-0779/2020\|EPI_ISL_508945\|2020-0 | ···················· | ···················· | ···················· | ···················· | ···················· | ···················· | |
| F | 3494 | hCoV-19/France/ARA-08067/2020\|EPI_ISL_508946\|2020- | ···················· | ···················· | ···················· | ···················· | ···················· | ···················· | |
| F | 3495 | hCoV-19/France/ARA-0814/2020\|EPI_ISL_508947\|2020-0 | ···················· | ···················· | ···················· | ···················· | ···················· | ···················· | |
| F | 3496 | hCoV-19/France/ARA-08417/2020\|EPI_ISL_508948\|2020- | ···················· | ···················· | ···················· | ···················· | ···················· | ···················· | |
| F | 3497 | hCoV-19/France/ARA-12155/2020\|EPI_ISL_508949\|2020- | ···················· | ···················· | ···················· | ···················· | ···················· | ···················· | |

| | | | | | | | | | |
|---|---|---|---|---|---|---|---|---|---|
| F | 3498 | hCoV-19/France/ARA-12282/2020\|EPI_ISL_508950\|2020- | ···················· | ···················· | ···················· | ···················· | ···················· | ···················· | |
| F | 3499 | hCoV-19/France/ARA-12287/2020\|EPI_ISL_508951\|2020- | ···················· | ···················· | ···················· | ···················· | ···················· | ···················· | |
| F | 3500 | hCoV-19/France/ARA-12638/2020\|EPI_ISL_508952\|2020- | ···················· | ···················· | ···················· | ···················· | ···················· | ···················· | |
| F | 3501 | hCoV-19/France/ARA-12742/2020\|EPI_ISL_508953\|2020- | ···················· | ···················· | ···················· | ···················· | ···················· | ···················· | |
| F | 3502 | hCoV-19/France/ARA-13353/2020\|EPI_ISL_508954\|2020- | ···················· | ···················· | ···················· | ···················· | ···················· | ···················· | |
| F | 3503 | hCoV-19/France/ARA-13367/2020\|EPI_ISL_508955\|2020- | ···················· | ···················· | ···················· | ···················· | ···················· | ···················· | |
| F | 3504 | hCoV-19/France/ARA-13374/2020\|EPI_ISL_508956\|2020- | ···················· | ···················· | ···················· | ···················· | ···················· | ···················· | |
| F | 3505 | hCoV-19/France/ARA-13515/2020\|EPI_ISL_508957\|2020- | ···················· | ···················· | ···················· | ···················· | ···················· | ···················· | |
| F | 3506 | hCoV-19/France/ARA-17177/2020\|EPI_ISL_508958\|2020- | ···················· | ···················· | ···················· | ···················· | ···················· | ···················· | |
| F | 3507 | hCoV-19/France/ARA-17330/2020\|EPI_ISL_508959\|2020- | ···················· | ···················· | ···················· | ···················· | ···················· | ···················· | |
| F | 3508 | hCoV-19/France/ARA-17747/2020\|EPI_ISL_508960\|2020- | ···················· | ···················· | ···················· | ···················· | ···················· | ···················· | |
| F | 3509 | hCoV-19/France/ARA-1783/2020\|EPI_ISL_508961\|2020-0 | ···················· | ···················· | ···················· | ···················· | ···················· | ···················· | |
| F | 3510 | hCoV-19/France/ARA-1811/2020\|EPI_ISL_508962\|2020-0 | ···················· | ···················· | ···················· | ···················· | ···················· | ···················· | |
| F | 3511 | hCoV-19/France/ARA-18272/2020\|EPI_ISL_508964\|2020- | ···················· | ···················· | ···················· | ···················· | ···················· | ···················· | |
| F | 3512 | hCoV-19/France/ARA-18275/2020\|EPI_ISL_508965\|2020- | ···················· | ···················· | ···················· | ···················· | ···················· | ···················· | |
| F | 3513 | hCoV-19/France/ARA-18314/2020\|EPI_ISL_508966\|2020- | ···················· | ···················· | ···················· | ···················· | ···················· | ···················· | |
| F | 3514 | hCoV-19/France/ARA-18366/2020\|EPI_ISL_508967\|2020- | ···················· | ···················· | ···················· | ···················· | ···················· | ···················· | |
| F | 3515 | hCoV-19/France/ARA-18437/2020\|EPI_ISL_508968\|2020- | ···················· | ···················· | ···················· | ···················· | ···················· | ···················· | |
| F | 3516 | hCoV-19/France/ARA-18440/2020\|EPI_ISL_508969\|2020- | ···················· | ···················· | ···················· | ···················· | ···················· | ···················· | |
| F | 3517 | hCoV-19/USA/CA-ALSR-1740/2020\|EPI_ISL_512161\|2020- | ···················· | ·A·················· | ···················· | ···················· | ···················· | ···················· | |
| F | 3518 | hCoV-19/USA/CA-ALSR-1743/2020\|EPI_ISL_512162\|2020- | ···················· | ·A·················· | ···················· | ···················· | ···················· | ···················· | |
| F | 3519 | hCoV-19/USA/CA-ALSR-1749/2020\|EPI_ISL_512164\|2020- | ···················· | ·A·················· | ···················· | ···················· | ···················· | ···················· | |
| F | 3520 | hCoV-19/USA/CA-ALSR-1761/2020\|EPI_ISL_512170\|2020- | ···················· | ·A·················· | ···················· | ···················· | ···················· | ···················· | |
| F | 3521 | hCoV-19/USA/CA-ALSR-1763/2020\|EPI_ISL_512171\|2020- | ···················· | ·A·················· | ···················· | ···················· | ···················· | ···················· | |
| F | 3522 | hCoV-19/USA/CA-ALSR-1771/2020\|EPI_ISL_512172\|2020- | ···················· | ·A·················· | ···················· | ···················· | ···················· | ···················· | |
| F | 3523 | hCoV-19/USA/CA-ALSR-1772/2020\|EPI_ISL_512173\|2020- | ···················· | ···················· | ···················· | ···················· | ···················· | ···················· | |
| F | 3524 | hCoV-19/USA/CA-ALSR-1773/2020\|EPI_ISL_512174\|2020- | ···················· | ·A·················· | ···················· | ···················· | ···················· | ···················· | |
| F | 3525 | hCoV-19/USA/CA-ALSR-1775/2020\|EPI_ISL_512175\|2020- | ···················· | ·A·················· | ···················· | ···················· | ···················· | ···················· | |
| F | 3526 | hCoV-19/USA/CA-ALSR-1776/2020\|EPI_ISL_512176\|2020- | ···················· | ·A·················· | ···················· | ···················· | ···················· | ···················· | |
| F | 3527 | hCoV-19/USA/CA-ALSR-1779/2020\|EPI_ISL_512178\|2020- | ···················· | ·A·················· | ···················· | ···················· | ···················· | ···················· | |
| F | 3528 | hCoV-19/USA/CA-ALSR-1781/2020\|EPI_ISL_512179\|2020- | ···················· | ···················· | ···················· | ···················· | ···················· | ···················· | |
| F | 3529 | hCoV-19/USA/CA-ALSR-1782/2020\|EPI_ISL_512180\|2020- | ···················· | ·A·················· | ···················· | ···················· | ···················· | ···················· | |
| F | 3530 | hCoV-19/USA/CA-ALSR-1783/2020\|EPI_ISL_512181\|2020- | ··················G· | ···················· | ···················· | ···················· | ···················· | ···················· | 1 |
| F | 3531 | hCoV-19/USA/CA-ALSR-1784/2020\|EPI_ISL_512182\|2020- | ···················· | ·A·················· | ···················· | ···················· | ···················· | ···················· | |
| F | 3532 | hCoV-19/USA/CA-ALSR-1785/2020\|EPI_ISL_512183\|2020- | ···················· | ·A·················· | ···················· | ···················· | ···················· | ···················· | |
| F | 3533 | hCoV-19/USA/CA-ALSR-1786/2020\|EPI_ISL_512184\|2020- | ···················· | ·A·················· | ···················· | ···················· | ···················· | ···················· | |
| F | 3534 | hCoV-19/USA/CA-ALSR-1788/2020\|EPI_ISL_512185\|2020- | ···················· | ·A·················· | ···················· | ···················· | ···················· | ···················· | |
| F | 3535 | hCoV-19/USA/CA-ALSR-1789/2020\|EPI_ISL_512186\|2020- | ···················· | ·A·················· | ···················· | ···················· | ···················· | ···················· | |
| F | 3536 | hCoV-19/USA/CA-ALSR-1791/2020\|EPI_ISL_512187\|2020- | ···················· | ·A·················· | ···················· | ···················· | ···················· | ···················· | |
| F | 3537 | hCoV-19/USA/CA-ALSR-1793/2020\|EPI_ISL_512188\|2020- | ···················· | ·A·················· | ···················· | ···················· | ···················· | ···················· | |
| F | 3538 | hCoV-19/USA/CA-ALSR-1795/2020\|EPI_ISL_512189\|2020- | ···················· | ·A·················· | ···················· | ···················· | ···················· | ···················· | |
| F | 3539 | hCoV-19/USA/CA-ALSR-1800/2020\|EPI_ISL_512191\|2020- | ···················· | ···················· | ···················· | ···················· | ···················· | ···················· | |
| F | 3540 | hCoV-19/USA/CA-ALSR-1801/2020\|EPI_ISL_512192\|2020- | ···················· | ·A·················· | ···················· | ···················· | ···················· | ···················· | |
| F | 3541 | hCoV-19/USA/CA-ALSR-1802/2020\|EPI_ISL_512193\|2020- | ···················· | ·A·················· | ···················· | ···················· | ···················· | ···················· | |
| F | 3542 | hCoV-19/USA/CA-ALSR-1804/2020\|EPI_ISL_512194\|2020- | ···················· | ···················· | ···················· | ···················· | ···················· | ···················· | |
| F | 3543 | hCoV-19/USA/CA-ALSR-1816/2020\|EPI_ISL_512195\|2020- | ···················· | ·A·················· | ···················· | ···················· | ···················· | ···················· | |
| F | 3544 | hCoV-19/USA/CA-ALSR-1818/2020\|EPI_ISL_512196\|2020- | ···················· | ·A·················· | ···················· | ···················· | ···················· | ···················· | |
| F | 3545 | hCoV-19/USA/CA-ALSR-1826/2020\|EPI_ISL_512197\|2020- | ···················· | ·A·················· | ···················· | ···················· | ···················· | ···················· | |
| F | 3546 | hCoV-19/USA/CA-ALSR-1827/2020\|EPI_ISL_512198\|2020- | ···················· | ·A·················· | ···················· | ···················· | ···················· | ···················· | |
| F | 3547 | hCoV-19/USA/CA-ALSR-1829/2020\|EPI_ISL_512199\|2020- | ···················· | ···················· | ···················· | ···················· | ···················· | ···················· | |
| F | 3548 | hCoV-19/Australia/NSW758/2020\|EPI_ISL_513400\|2020- | ···················· | ···················· | ···················· | ···················· | ···················· | ···················· | |
| F | 3549 | hCoV-19/Australia/NSW759/2020\|EPI_ISL_513401\|2020- | ···················· | ···················· | ···················· | ···················· | ···················· | ···················· | |
| F | 3550 | hCoV-19/Australia/NSW760/2020\|EPI_ISL_513402\|2020- | ···················· | ···················· | ···················· | ···················· | ···················· | ···················· | |
| F | 3551 | hCoV-19/Australia/NSW761/2020\|EPI_ISL_513403\|2020- | ···················· | ···················· | ···················· | ···················· | ···················· | ···················· | |
| F | 3552 | hCoV-19/Australia/NSW762/2020\|EPI_ISL_513404\|2020- | ···················· | ···················· | ···················· | ···················· | ···················· | ···················· | |
| F | 3553 | hCoV-19/Australia/NSW763/2020\|EPI_ISL_513405\|2020- | ···················· | ···················· | ···················· | ···················· | ···················· | ···················· | |
| F | 3554 | hCoV-19/Australia/NSW764/2020\|EPI_ISL_513406\|2020- | ···················· | ···················· | ···················· | ···················· | ···················· | ···················· | |
| F | 3555 | hCoV-19/Australia/NSW765/2020\|EPI_ISL_513407\|2020- | ···················· | ···················· | ···················· | ···················· | ···················· | ···················· | |
| F | 3556 | hCoV-19/Australia/NSW766/2020\|EPI_ISL_513408\|2020- | ···················· | ···················· | ···················· | ···················· | ···················· | ···················· | |
| F | 3557 | hCoV-19/Australia/NSW767/2020\|EPI_ISL_513409\|2020- | ···················· | ···················· | ···················· | ···················· | ···················· | ···················· | |
| F | 3558 | hCoV-19/Australia/NSW768/2020\|EPI_ISL_513410\|2020- | ···················· | ···················· | ···················· | ···················· | ···················· | ···················· | |
| F | 3559 | hCoV-19/Australia/NSW769/2020\|EPI_ISL_513411\|2020- | ···················· | ···················· | ···················· | ···················· | ···················· | ···················· | |
| F | 3560 | hCoV-19/Australia/NSW770/2020\|EPI_ISL_513412\|2020- | ···················· | ···················· | ···················· | ···················· | ···················· | ···················· | |
| F | 3561 | hCoV-19/Australia/NSW771/2020\|EPI_ISL_513413\|2020- | ···················· | ···················· | ···················· | ···················· | ···················· | ···················· | |
| F | 3562 | hCoV-19/USA/ME-HETL-J0001/2020\|EPI_ISL_513414\|2020 | ···················· | ···················· | ···················· | ···················· | ···················· | ···················· | |
| F | 3563 | hCoV-19/USA/ME-HETL-J0002/2020\|EPI_ISL_513415\|2020 | ···················· | ···················· | ···················· | ···················· | ···················· | ···················· | |
| F | 3564 | hCoV-19/USA/ME-HETL-J0003/2020\|EPI_ISL_513416\|2020 | ···················· | ···················· | ···················· | ···················· | ···················· | ···················· | |
| F | 3565 | hCoV-19/USA/ME-HETL-J0005/2020\|EPI_ISL_513418\|2020 | ···················· | ···················· | ···················· | ···················· | ···················· | ···················· | |
| F | 3566 | hCoV-19/USA/ME-HETL-J0006/2020\|EPI_ISL_513419\|2020 | ···················· | ···················· | ···················· | ···················· | ···················· | ···················· | |
| F | 3567 | hCoV-19/USA/ME-HETL-J0007/2020\|EPI_ISL_513420\|2020 | ···················· | ···················· | ···················· | ···················· | ···················· | ···················· | |
| F | 3568 | hCoV-19/USA/ME-HETL-J0008/2020\|EPI_ISL_513421\|2020 | ···················· | ···················· | ···················· | ···················· | ···················· | ···················· | |
| F | 3569 | hCoV-19/USA/ME-HETL-J0009/2020\|EPI_ISL_513422\|2020 | ···················· | ···················· | ···················· | ···················· | ···················· | ···················· | |
| F | 3570 | hCoV-19/USA/ME-HETL-J0010/2020\|EPI_ISL_513423\|2020 | ···················· | ···················· | ···················· | ···················· | ···················· | ···················· | |
| F | 3571 | hCoV-19/USA/ME-HETL-J0011/2020\|EPI_ISL_513424\|2020 | ···················· | ···················· | ···················· | ···················· | ···················· | ···················· | |
| F | 3572 | hCoV-19/USA/ME-HETL-J0012/2020\|EPI_ISL_513425\|2020 | ···················· | ···················· | ···················· | ···················· | ···················· | ···················· | |
| F | 3573 | hCoV-19/USA/ME-HETL-J0013/2020\|EPI_ISL_513426\|2020 | ···················· | ···················· | ···················· | ···················· | ···················· | ···················· | |
| F | 3574 | hCoV-19/USA/ME-HETL-J0081/2020\|EPI_ISL_513492\|2020 | ···················· | ···················· | ···················· | ···················· | ···················· | ···················· | |
| F | 3575 | hCoV-19/USA/ME-HETL-J0083/2020\|EPI_ISL_513493\|2020 | ···················· | ···················· | ···················· | ···················· | ···················· | ···················· | |
| F | 3576 | hCoV-19/USA/ME-HETL-J0084/2020\|EPI_ISL_513494\|2020 | ···················· | ···················· | ···················· | ···················· | ···················· | ···················· | |
| F | 3577 | hCoV-19/USA/ME-HETL-J0085/2020\|EPI_ISL_513495\|2020 | ···················· | ···················· | ···················· | ···················· | ···················· | ···················· | |

| | | | | | | | |
|---|---|---|---|---|---|---|---|
| F | 3578 hCoV-19/Russia/MOW-PMU-0417/2020\|EPI_ISL_514750\|20 | .................... | .................... | .................... | .................... | .................... | .................... |
| F | 3579 hCoV-19/Italy/CAM-CEINGE-S66/2020\|EPI_ISL_514751\|2 | .................... | .................... | .................... | .................... | .................... | .................... |
| F | 3580 hCoV-19/Iran/IRVSH-2/2020\|EPI_ISL_514753\|2020-05-0 | .................... | .A.................. | .................... | .................... | .................... | .................... |
| F | 3581 hCoV-19/South Korea/KCDC2353/2020\|EPI_ISL_514756\|2 | .................... | .................... | .................... | .................... | .................... | .................... |
| F | 3582 hCoV-19/South Korea/KCDC2354/2020\|EPI_ISL_514757\|2 | .................... | .................... | .................... | .................... | .................... | .................... |
| F | 3583 hCoV-19/Germany/BY-UHR-25568233R/2020\|EPI_ISL_5254 | .................... | .................... | .................... | .................... | .................... | .................... |
| F | 3584 hCoV-19/Germany/BY-UHR-25578139L/2020\|EPI_ISL_5254 | .................... | .................... | .................... | .................... | .................... | .................... |
| F | 3585 hCoV-19/DRC/5240/2020\|EPI_ISL_496492\|2020-05-03 | .................... | .................... | .................... | .................... | .................... | .................... |
| F | 3586 hCoV-19/DRC/5913/2020\|EPI_ISL_496497\|2020-05-07 | .................... | .................... | .................... | .................... | .................... | .................... |
| F | 3587 hCoV-19/DRC/5927/2020\|EPI_ISL_496498\|2020-05-07 | .................... | .................... | .................... | .................... | .................... | .................... |
| F | 3588 hCoV-19/DRC/5957/2020\|EPI_ISL_496499\|2020-05-07 | .................... | .................... | .................... | .................... | .................... | .................... |
| F | 3589 hCoV-19/Hong Kong/HKU-200723-008/2020\|EPI_ISL_4977 | .................... | .................... | .................... | .................... | .................... | .................... |
| F | 3590 hCoV-19/Hong Kong/HKU-200723-009/2020\|EPI_ISL_4977 | .................... | .................... | .................... | .................... | .................... | .................... |
| F | 3591 hCoV-19/Hong Kong/HKU-200723-010/2020\|EPI_ISL_4977 | .................... | .................... | .................... | .................... | .................... | .................... |
| F | 3592 hCoV-19/Hong Kong/HKU-200723-011/2020\|EPI_ISL_4977 | .................... | .................... | .................... | .................... | .................... | .................... |
| F | 3593 hCoV-19/Hong Kong/HKU-200723-013/2020\|EPI_ISL_4977 | .................... | .................... | .................... | .................... | .................... | .................... |
| F | 3594 hCoV-19/England/LIVE-A2774/2020\|EPI_ISL_500007\|202 | .................... | .................... | .................... | .................... | .................... | .................... |
| F | 3595 hCoV-19/England/LIVE-A27FC/2020\|EPI_ISL_500008\|202 | .................... | .................... | .................... | .................... | .................... | .................... |
| F | 3596 hCoV-19/England/LIVE-A2853/2020\|EPI_ISL_500009\|202 | .................... | .................... | .................... | .................... | .................... | .................... |
| F | 3597 hCoV-19/England/LIVE-A2880/2020\|EPI_ISL_500010\|202 | .................... | .................... | .................... | .................... | .................... | .................... |
| F | 3599 hCoV-19/England/LIVE-A296F/2020\|EPI_ISL_500012\|202 | .................... | .................... | .................... | .................... | .................... | .................... |
| F | 3600 hCoV-19/England/LIVE-A298D/2020\|EPI_ISL_500013\|202 | .................... | .................... | .................... | .................... | .................... | .................... |
| F | 3603 hCoV-19/England/LIVE-A2EE8/2020\|EPI_ISL_500016\|202 | .................... | .................... | .................... | .................... | .................... | .................... |
| F | 3604 hCoV-19/England/LIVE-A3C47/2020\|EPI_ISL_500017\|202 | .................... | .................... | .................... | .................... | .................... | .................... |
| F | 3606 hCoV-19/England/LIVE-A3C74/2020\|EPI_ISL_500019\|202 | .................... | .................... | .................... | .................... | .................... | .................... |
| F | 3607 hCoV-19/England/LIVE-A3C92/2020\|EPI_ISL_500020\|202 | .................... | .................... | .................... | .................... | .................... | .................... |
| F | 3608 hCoV-19/England/LIVE-A3CA1/2020\|EPI_ISL_500021\|202 | .................... | .................... | .................... | .................... | .................... | .................... |
| F | 3609 hCoV-19/England/LIVE-A3CDE/2020\|EPI_ISL_500022\|202 | .................... | .................... | .................... | .................... | .................... | .................... |
| F | 3610 hCoV-19/England/LIVE-A3D35/2020\|EPI_ISL_500023\|202 | .................... | .................... | .................... | .................... | .................... | .................... |
| F | 3611 hCoV-19/England/LIVE-A3D44/2020\|EPI_ISL_500024\|202 | .................... | .................... | .................... | .................... | .................... | .................... |
| F | 3613 hCoV-19/England/LIVE-A3D9F/2020\|EPI_ISL_500026\|202 | .................... | .................... | .................... | .................... | .................... | .................... |
| F | 3614 hCoV-19/England/LIVE-A3DAE/2020\|EPI_ISL_500027\|202 | .................... | .................... | .................... | .................... | .................... | .................... |
| F | 3615 hCoV-19/England/LIVE-A3E32/2020\|EPI_ISL_500028\|202 | .................... | .................... | .................... | .................... | .................... | .................... |
| F | 3616 hCoV-19/England/LIVE-A4499/2020\|EPI_ISL_500029\|202 | .................... | .................... | .................... | .................... | .................... | .................... |
| F | 3618 hCoV-19/England/LIVE-A44F3/2020\|EPI_ISL_500031\|202 | .................... | .................... | .................... | .................... | .................... | .................... |
| F | 3624 hCoV-19/England/LIVE-A4736/2020\|EPI_ISL_500037\|202 | .................... | .................... | .................... | .................... | .................... | .................... |
| F | 3625 hCoV-19/England/LIVE-A4745/2020\|EPI_ISL_500038\|202 | .................... | .................... | .................... | .................... | .................... | .................... |
| F | 3627 hCoV-19/England/LIVE-A4763/2020\|EPI_ISL_500040\|202 | .................... | .................... | .................... | .................... | .................... | .................... |
| F | 3629 hCoV-19/England/LIVE-A47AF/2020\|EPI_ISL_500042\|202 | .................... | .................... | .................... | .................... | .................... | .................... |
| F | 3631 hCoV-19/England/LIVE-A47EB/2020\|EPI_ISL_500044\|202 | .................... | .................... | .................... | .................... | .................... | .................... |
| F | 3635 hCoV-19/England/LIVE-A4833/2020\|EPI_ISL_500048\|202 | .................... | .................... | .................... | .................... | .................... | .................... |
| F | 3638 hCoV-19/USA/MI-MDHHS-SC21428/2020\|EPI_ISL_507682\|2 | .................... | .................... | .................... | .................... | .................... | .................... |
| F | 3639 hCoV-19/USA/MI-MDHHS-SC21430/2020\|EPI_ISL_507684\|2 | .................... | .................... | .................... | .................... | .................... | .................... |
| F | 3640 hCoV-19/USA/MI-MDHHS-SC21431/2020\|EPI_ISL_507685\|2 | .................... | .................... | .................... | .................... | .................... | .................... |
| F | 3641 hCoV-19/USA/MI-MDHHS-SC21432/2020\|EPI_ISL_507686\|2 | .................... | .................... | .................... | .................... | .................... | .................... |
| F | 3642 hCoV-19/USA/MI-MDHHS-SC21434/2020\|EPI_ISL_507688\|2 | .................... | .................... | .................... | .................... | .................... | .................... |
| F | 3643 hCoV-19/USA/MI-MDHHS-SC21435/2020\|EPI_ISL_507689\|2 | .................... | .................... | .................... | .................... | .................... | .................... |
| F | 3644 hCoV-19/USA/MI-MDHHS-SC21436/2020\|EPI_ISL_507690\|2 | .................... | .................... | .................... | .................... | .................... | .................... |
| F | 3645 hCoV-19/USA/MI-MDHHS-SC21437/2020\|EPI_ISL_507691\|2 | .................... | .................... | .................... | .................... | .................... | .................... |
| F | 3646 hCoV-19/USA/MI-MDHHS-SC21438/2020\|EPI_ISL_507692\|2 | .................... | .................... | .................... | .................... | .................... | .................... |
| F | 3647 hCoV-19/USA/MI-MDHHS-SC21439/2020\|EPI_ISL_507693\|2 | .................... | .................... | .................... | .................... | .................... | .................... |
| F | 3648 hCoV-19/USA/MI-MDHHS-SC21440/2020\|EPI_ISL_507694\|2 | .................... | .................... | .................... | .................... | .................... | .................... |
| F | 3649 hCoV-19/USA/MI-MDHHS-SC21441/2020\|EPI_ISL_507695\|2 | .................... | .................... | .................... | .................... | .................... | .................... |
| F | 3650 hCoV-19/USA/MI-MDHHS-SC21442/2020\|EPI_ISL_507696\|2 | .................... | .................... | .................... | .................... | .................... | .................... |
| F | 3651 hCoV-19/USA/MI-MDHHS-SC21443/2020\|EPI_ISL_507697\|2 | .................... | .................... | .................... | .................... | .................... | .................... |
| F | 3652 hCoV-19/USA/MI-MDHHS-SC21444/2020\|EPI_ISL_507698\|2 | .................... | .................... | .................... | .................... | .................... | .................... |
| F | 3653 hCoV-19/USA/MI-MDHHS-SC21445/2020\|EPI_ISL_507699\|2 | .................... | .................... | .................... | .................... | .................... | .................... |
| F | 3654 hCoV-19/France/ARA-15119/2020\|EPI_ISL_508900\|2020- | .................... | .................... | .................... | .................... | .................... | .................... |
| F | 3655 hCoV-19/France/ARA-15126/2020\|EPI_ISL_508901\|2020- | .................... | .................... | .................... | .................... | .................... | .................... |
| F | 3656 hCoV-19/France/ARA-15224/2020\|EPI_ISL_508902\|2020- | .................... | .................... | .................... | .................... | .................... | .................... |
| F | 3657 hCoV-19/France/ARA-15296/2020\|EPI_ISL_508903\|2020- | .................... | .................... | .................... | .................... | .................... | .................... |
| F | 3658 hCoV-19/France/ARA-16170/2020\|EPI_ISL_508904\|2020- | .................... | .................... | .................... | .................... | .................... | .................... |
| F | 3659 hCoV-19/France/ARA-16213/2020\|EPI_ISL_508905\|2020- | .................... | .................... | .................... | .................... | .................... | .................... |
| F | 3660 hCoV-19/France/ARA-18487/2020\|EPI_ISL_508970\|2020- | .................... | .................... | .................... | .................... | .................... | .................... |
| F | 3661 hCoV-19/France/ARA-18634/2020\|EPI_ISL_508972\|2020- | .................... | .................... | .................... | .................... | .................... | .................... |
| F | 3662 hCoV-19/France/ARA-18748/2020\|EPI_ISL_508973\|2020- | .................... | .................... | .................... | .................... | .................... | .................... |
| F | 3663 hCoV-19/France/ARA-18783/2020\|EPI_ISL_508974\|2020- | .................... | .................... | .................... | .................... | .................... | .................... |
| F | 3664 hCoV-19/France/ARA-18826/2020\|EPI_ISL_508975\|2020- | .................... | .................... | .................... | .................... | .................... | .................... |
| F | 3665 hCoV-19/France/ARA-19119/2020\|EPI_ISL_508976\|2020- | .................... | .................... | .................... | .................... | .................... | .................... |
| F | 3666 hCoV-19/France/ARA-19311/2020\|EPI_ISL_508977\|2020- | .................... | .................... | .................... | .................... | .................... | .................... |
| F | 3667 hCoV-19/France/ARA-19340/2020\|EPI_ISL_508978\|2020- | .................... | .................... | .................... | .................... | .................... | .................... |
| F | 3668 hCoV-19/France/ARA-19556/2020\|EPI_ISL_508979\|2020- | .................... | .................... | .................... | .................... | .................... | .................... |
| F | 3669 hCoV-19/France/ARA-19570/2020\|EPI_ISL_508980\|2020- | .................... | .................... | .................... | .................... | .................... | .................... |
| F | 3670 hCoV-19/France/ARA-19928/2020\|EPI_ISL_508981\|2020- | .................... | .................... | .................... | .................... | .................... | .................... |
| F | 3671 hCoV-19/France/ARA-20073/2020\|EPI_ISL_508982\|2020- | .................... | .................... | .................... | .................... | .................... | .................... |
| F | 3672 hCoV-19/France/ARA-20154/2020\|EPI_ISL_508983\|2020- | .................... | .................... | .................... | .................... | .................... | .................... |
| F | 3673 hCoV-19/France/ARA-20193/2020\|EPI_ISL_508984\|2020- | .................... | .................... | .................... | .................... | .................... | .................... |
| F | 3674 hCoV-19/France/ARA-20254/2020\|EPI_ISL_508985\|2020- | .................... | .................... | .................... | .................... | .................... | .................... |
| F | 3675 hCoV-19/France/ARA-13746/2020\|EPI_ISL_508986\|2020- | .................... | .................... | .................... | .................... | .................... | .................... |
| F | 3676 hCoV-19/France/ARA-16665/2020\|EPI_ISL_508987\|2020- | .................... | .................... | .................... | .................... | .................... | .................... |

| | | | | | | | | | |
|---|---|---|---|---|---|---|---|---|---|
| F | 3677 | hCoV-19/France/ARA-20869/2020\|EPI_ISL_508990\|2020- | ···················· | ···················· | ···················· | ···················· | ···················· | ···················· | |
| F | 3678 | hCoV-19/France/ARA-21154/2020\|EPI_ISL_508991\|2020- | ···················· | ···················· | ···················· | ···················· | ···················· | ···················· | |
| F | 3679 | hCoV-19/France/ARA-21210/2020\|EPI_ISL_508993\|2020- | ···················· | ···················· | ···················· | ···················· | ···················· | ···················· | |
| F | 3680 | hCoV-19/France/ARA-21399/2020\|EPI_ISL_508994\|2020- | ···················· | ···················· | ···················· | ···················· | ···················· | ···················· | |
| F | 3681 | hCoV-19/France/ARA-21941/2020\|EPI_ISL_508995\|2020- | ···················· | ···················· | ···················· | ···················· | ···················· | ···················· | |
| F | 3682 | hCoV-19/France/ARA-22088/2020\|EPI_ISL_508996\|2020- | ···················· | ···················· | ···················· | ···················· | ···················· | ···················· | |
| F | 3683 | hCoV-19/France/ARA-22132/2020\|EPI_ISL_508997\|2020- | ···················· | ···················· | ···················· | ···················· | ···················· | ···················· | |
| F | 3684 | hCoV-19/France/ARA-10441/2020\|EPI_ISL_508998\|2020- | ···················· | ···················· | ···················· | ···················· | ···················· | ···················· | |
| F | 3685 | hCoV-19/France/ARA-10580/2020\|EPI_ISL_508999\|2020- | ···················· | ···················· | ···················· | ···················· | ···················· | ···················· | |
| F | 3686 | hCoV-19/Switzerland/ZH-ETHZ-220016/2020\|EPI_ISL_51 | ···················· | ···················· | ···················· | ···················· | ···················· | ···················· | |
| F | 3687 | hCoV-19/Switzerland/ZH-ETHZ-220018/2020\|EPI_ISL_51 | ···················· | ···················· | ···················· | ···················· | ···················· | ···················· | |
| F | 3688 | hCoV-19/Switzerland/SG-ETHZ-220019/2020\|EPI_ISL_51 | ···················· | ···················· | ···················· | ···················· | ···················· | ···················· | |
| F | 3689 | hCoV-19/Switzerland/SZ-ETHZ-220020/2020\|EPI_ISL_51 | ···················· | ···················· | ···················· | ···················· | ···················· | ···················· | |
| F | 3690 | hCoV-19/Switzerland/UR-ETHZ-220021/2020\|EPI_ISL_51 | ···················· | ·A·················· | ···················· | ···················· | ···················· | ···················· | |
| F | 3691 | hCoV-19/Switzerland/VD-ETHZ-220022/2020\|EPI_ISL_51 | ···················· | ···················· | ···················· | ···················· | ···················· | ···················· | |
| F | 3692 | hCoV-19/Switzerland/ZH-ETHZ-220023/2020\|EPI_ISL_51 | ···················· | ···················· | ···················· | ···················· | ···················· | ···················· | |
| F | 3693 | hCoV-19/Switzerland/BE-ETHZ-220026/2020\|EPI_ISL_51 | ···················· | ·········R·········· | ···················· | ···················· | ···················· | ···················· | |
| F | 3694 | hCoV-19/Switzerland/SH-ETHZ-220027/2020\|EPI_ISL_51 | ···················· | ···················· | ···················· | ···················· | ···················· | ···················· | |
| F | 3695 | hCoV-19/Switzerland/LU-ETHZ-220028/2020\|EPI_ISL_51 | ···················· | ···················· | ···················· | ···················· | ···················· | ···················· | |
| F | 3696 | hCoV-19/Switzerland/ZH-ETHZ-220029/2020\|EPI_ISL_51 | ···················· | ···················· | ···················· | ···················· | ···················· | ···················· | |
| F | 3697 | hCoV-19/Switzerland/GE-ETHZ-220030/2020\|EPI_ISL_51 | ···················· | ···················· | ···················· | ···················· | ···················· | ···················· | |
| F | 3698 | hCoV-19/Switzerland/AG-ETHZ-220032/2020\|EPI_ISL_51 | ···················· | ···················· | ···················· | ···················· | ···················· | ···················· | |
| F | 3699 | hCoV-19/Switzerland/BE-ETHZ-220034/2020\|EPI_ISL_51 | ···················· | ···················· | ···················· | ···················· | ···················· | ·················S·· | 1 |
| F | 3700 | hCoV-19/Switzerland/VD-ETHZ-220035/2020\|EPI_ISL_51 | ···················· | ···················· | ···················· | ···················· | ···················· | ···················· | |
| F | 3701 | hCoV-19/Switzerland/BL-ETHZ-220036/2020\|EPI_ISL_51 | ···················· | ···················· | ···················· | ···················· | ···················· | ···················· | |
| F | 3702 | hCoV-19/Switzerland/BE-ETHZ-220037/2020\|EPI_ISL_51 | ···················· | ···················· | ···················· | ···················· | ···················· | ···················· | |
| F | 3703 | hCoV-19/Switzerland/ZH-ETHZ-220040/2020\|EPI_ISL_51 | ···················· | ···················· | ···················· | ···················· | ···················· | ···················· | |
| F | 3704 | hCoV-19/Switzerland/BL-ETHZ-220041/2020\|EPI_ISL_51 | ···················· | ···················· | ···················· | ···················· | ···················· | ···················· | |
| F | 3705 | hCoV-19/Switzerland/SG-ETHZ-220043/2020\|EPI_ISL_51 | ···················· | ···················· | ···················· | ···················· | ···················· | ···················· | |
| F | 3706 | hCoV-19/Switzerland/BL-ETHZ-220046/2020\|EPI_ISL_51 | ···················· | ···················· | ···················· | ···················· | ···················· | ···················· | |
| F | 3707 | hCoV-19/Switzerland/ZH-ETHZ-220047/2020\|EPI_ISL_51 | ···················· | ···················· | ···················· | ···················· | ···················· | ···················· | |
| F | 3708 | hCoV-19/Switzerland/ZH-ETHZ-220057/2020\|EPI_ISL_51 | ···················· | ···················· | ···················· | ···················· | ···················· | ···················· | |
| F | 3709 | hCoV-19/Switzerland/BE-ETHZ-220058/2020\|EPI_ISL_51 | ···················· | ···················· | ···················· | ···················· | ···················· | ···················· | |
| F | 3710 | hCoV-19/Switzerland/BE-ETHZ-220059/2020\|EPI_ISL_51 | ···················· | ·A·················· | ···················· | ···················· | ···················· | ···················· | |
| F | 3711 | hCoV-19/USA/AK-PHL621/2020\|EPI_ISL_512156\|2020-06- | ···················· | ···················· | ···················· | ···················· | ···················· | ···················· | |
| F | 3712 | hCoV-19/USA/AK-PHL622/2020\|EPI_ISL_512157\|2020-06- | ···················· | ···················· | ···················· | ···················· | ···················· | ···················· | |
| F | 3713 | hCoV-19/USA/CA-ALSR-1735/2020\|EPI_ISL_512158\|2020- | ···················· | ·A·················· | ···················· | ···················· | ···················· | ···················· | |
| F | 3714 | hCoV-19/USA/CA-ALSR-1739/2020\|EPI_ISL_512160\|2020- | ···················· | ···················· | ···················· | ···················· | ···················· | ···················· | |
| F | 3715 | hCoV-19/USA/CA-ALSR-1747/2020\|EPI_ISL_512163\|2020- | ···················· | ···················· | ···················· | ···················· | ···················· | ···················· | |
| F | 3716 | hCoV-19/USA/CA-ALSR-1751/2020\|EPI_ISL_512165\|2020- | ···················· | ·A·················· | ···················· | ···················· | ···················· | ···················· | |
| F | 3717 | hCoV-19/USA/CA-ALSR-1753/2020\|EPI_ISL_512166\|2020- | ···················· | ·A·················· | ···················· | ···················· | ···················· | ···················· | |
| F | 3718 | hCoV-19/USA/CA-ALSR-1756/2020\|EPI_ISL_512167\|2020- | ···················· | ·A·················· | ···················· | ···················· | ···················· | ···················· | |
| F | 3719 | hCoV-19/USA/CA-ALSR-1759/2020\|EPI_ISL_512168\|2020- | ···················· | ···················· | ···················· | ···················· | ···················· | ···················· | |
| F | 3720 | hCoV-19/USA/CA-ALSR-1760/2020\|EPI_ISL_512169\|2020- | ···················· | ·A·················· | ···················· | ···················· | ···················· | ···················· | |
| F | 3721 | hCoV-19/Czech Republic/NRL_1259/2020\|EPI_ISL_51331 | ···················· | ···················· | ···················· | ···················· | ···················· | ···················· | |
| F | 3722 | hCoV-19/Australia/NSW674/2020\|EPI_ISL_513317\|2020- | ···················· | ···················· | ···················· | ···················· | ···················· | ···················· | |
| F | 3723 | hCoV-19/Australia/NSW675/2020\|EPI_ISL_513318\|2020- | ···················· | ···················· | ···················· | ···················· | ···················· | ···················· | |
| F | 3724 | hCoV-19/Australia/NSW676/2020\|EPI_ISL_513319\|2020- | ···················· | ···················· | ···················· | ···················· | ···················· | ···················· | |
| F | 3725 | hCoV-19/Australia/NSW677/2020\|EPI_ISL_513320\|2020- | ···················· | ···················· | ···················· | ···················· | ···················· | ···················· | |
| F | 3726 | hCoV-19/Australia/NSW678/2020\|EPI_ISL_513321\|2020- | ···················· | ···················· | ···················· | ···················· | ···················· | ···················· | |
| F | 3727 | hCoV-19/Australia/NSW679/2020\|EPI_ISL_513322\|2020- | ···················· | ···················· | ···················· | ···················· | ···················· | ···················· | |
| F | 3728 | hCoV-19/Australia/NSW680/2020\|EPI_ISL_513323\|2020- | ···················· | ···················· | ···················· | ···················· | ···················· | ···················· | |
| F | 3729 | hCoV-19/Australia/NSW681/2020\|EPI_ISL_513324\|2020- | ···················· | ···················· | ···················· | ···················· | ···················· | ···················· | |
| F | 3730 | hCoV-19/Australia/NSW682/2020\|EPI_ISL_513325\|2020- | ···················· | ···················· | ···················· | ···················· | ···················· | ···················· | |
| F | 3731 | hCoV-19/Australia/NSW683/2020\|EPI_ISL_513326\|2020- | ···················· | ···················· | ···················· | ···················· | ···················· | ···················· | |
| F | 3732 | hCoV-19/Australia/NSW684/2020\|EPI_ISL_513327\|2020- | ···················· | ···················· | ···················· | ···················· | ···················· | ···················· | |
| F | 3733 | hCoV-19/Australia/NSW685/2020\|EPI_ISL_513328\|2020- | ···················· | ···················· | ···················· | ···················· | ···················· | ···················· | |
| F | 3734 | hCoV-19/Australia/NSW686/2020\|EPI_ISL_513329\|2020- | ···················· | ···················· | ···················· | ···················· | ···················· | ···················· | |
| F | 3735 | hCoV-19/Australia/NSW687/2020\|EPI_ISL_513330\|2020- | ···················· | ···················· | ···················· | ···················· | ···················· | ···················· | |
| F | 3736 | hCoV-19/Australia/NSW688/2020\|EPI_ISL_513331\|2020- | ···················· | ···················· | ···················· | ···················· | ···················· | ···················· | |
| F | 3737 | hCoV-19/Australia/NSW689/2020\|EPI_ISL_513332\|2020- | ···················· | ···················· | ···················· | ···················· | ···················· | ···················· | |
| F | 3738 | hCoV-19/Australia/NSW690/2020\|EPI_ISL_513333\|2020- | ···················· | ···················· | ···················· | ···················· | ···················· | ···················· | |
| F | 3739 | hCoV-19/Australia/NSW691/2020\|EPI_ISL_513334\|2020- | ···················· | ···················· | ···················· | ···················· | ···················· | ···················· | |
| F | 3740 | hCoV-19/Australia/NSW692/2020\|EPI_ISL_513335\|2020- | ···················· | ···················· | ···················· | ···················· | ···················· | ···················· | |
| F | 3741 | hCoV-19/Australia/NSW693/2020\|EPI_ISL_513336\|2020- | ···················· | ···················· | ···················· | ···················· | ···················· | ···················· | |
| F | 3742 | hCoV-19/Australia/NSW694/2020\|EPI_ISL_513337\|2020- | ···················· | ···················· | ···················· | ···················· | ···················· | ···················· | |
| F | 3743 | hCoV-19/Australia/NSW695/2020\|EPI_ISL_513338\|2020- | ···················· | ···················· | ···················· | ···················· | ···················· | ···················· | |
| F | 3744 | hCoV-19/Australia/NSW696/2020\|EPI_ISL_513339\|2020- | ···················· | ···················· | ···················· | ···················· | ···················· | ···················· | |
| F | 3745 | hCoV-19/Australia/NSW697/2020\|EPI_ISL_513340\|2020- | ···················· | ···················· | ···················· | ···················· | ···················· | ···················· | |
| F | 3746 | hCoV-19/Australia/NSW698/2020\|EPI_ISL_513341\|2020- | ···················· | ···················· | ···················· | ···················· | ···················· | ···················· | |
| F | 3747 | hCoV-19/Australia/NSW699/2020\|EPI_ISL_513342\|2020- | ···················· | ···················· | ···················· | ···················· | ···················· | ···················· | |
| F | 3748 | hCoV-19/Australia/NSW700/2020\|EPI_ISL_513343\|2020- | ···················· | ···················· | ···················· | ···················· | ···················· | ···················· | |
| F | 3749 | hCoV-19/Australia/NSW701/2020\|EPI_ISL_513344\|2020- | ···················· | ···················· | ···················· | ···················· | ···················· | ···················· | |
| F | 3750 | hCoV-19/Australia/NSW702/2020\|EPI_ISL_513345\|2020- | ···················· | ···················· | ···················· | ···················· | ···················· | ···················· | |
| F | 3751 | hCoV-19/Australia/NSW703/2020\|EPI_ISL_513346\|2020- | ···················· | ···················· | ···················· | ···················· | ···················· | ···················· | |
| F | 3752 | hCoV-19/Australia/NSW704/2020\|EPI_ISL_513347\|2020- | ···················· | ···················· | ···················· | ···················· | ···················· | ···················· | |
| F | 3753 | hCoV-19/Australia/NSW705/2020\|EPI_ISL_513348\|2020- | ···················· | ···················· | ···················· | ···················· | ···················· | ···················· | |
| F | 3754 | hCoV-19/Australia/NSW706/2020\|EPI_ISL_513349\|2020- | ···················· | ···················· | ···················· | ···················· | ···················· | ···················· | |
| F | 3755 | hCoV-19/Australia/NSW707/2020\|EPI_ISL_513350\|2020- | ···················· | ···················· | ···················· | ···················· | ···················· | ···················· | |
| F | 3756 | hCoV-19/Australia/NSW708/2020\|EPI_ISL_513351\|2020- | ···················· | ···················· | ···················· | ···················· | ···················· | ···················· | |

| | | | | | | | | |
|---|---|---|---|---|---|---|---|---|
| F | 3757 | hCoV-19/Australia/NSW709/2020\|EPI_ISL_513352\|2020- | ···················· | ···················· | ···················· | ···················· | ···················· | ···················· |
| F | 3758 | hCoV-19/Australia/NSW710/2020\|EPI_ISL_513353\|2020- | ···················· | ···················· | ···················· | ···················· | ···················· | ···················· |
| F | 3759 | hCoV-19/Australia/NSW711/2020\|EPI_ISL_513354\|2020- | ···················· | ···················· | ···················· | ···················· | ···················· | ···················· |
| F | 3760 | hCoV-19/Australia/NSW712/2020\|EPI_ISL_513355\|2020- | ···················· | ···················· | ···················· | ···················· | ···················· | ···················· |
| F | 3761 | hCoV-19/Australia/NSW713/2020\|EPI_ISL_513356\|2020- | ···················· | ···················· | ···················· | ···················· | ···················· | ···················· |
| F | 3762 | hCoV-19/Australia/NSW714/2020\|EPI_ISL_513357\|2020- | ···················· | ···················· | ···················· | ···················· | ···················· | ···················· |
| F | 3763 | hCoV-19/Australia/NSW715/2020\|EPI_ISL_513358\|2020- | ···················· | ···················· | ···················· | ···················· | ···················· | ···················· |
| F | 3764 | hCoV-19/Australia/NSW716/2020\|EPI_ISL_513359\|2020- | ···················· | ···················· | ···················· | ···················· | ···················· | ···················· |
| F | 3765 | hCoV-19/USA/ME-HETL-J0069/2020\|EPI_ISL_513480\|2020 | ···················· | ···················· | ···················· | ···················· | ···················· | ···················· |
| F | 3766 | hCoV-19/USA/ME-HETL-J0070/2020\|EPI_ISL_513481\|2020 | ···················· | ···················· | ···················· | ···················· | ···················· | ···················· |
| F | 3767 | hCoV-19/USA/ME-HETL-J0071/2020\|EPI_ISL_513482\|2020 | ···················· | ···················· | ···················· | ···················· | ···················· | ···················· |
| F | 3768 | hCoV-19/USA/ME-HETL-J0072/2020\|EPI_ISL_513483\|2020 | ···················· | ···················· | ···················· | ···················· | ···················· | ···················· |
| F | 3769 | hCoV-19/USA/ME-HETL-J0073/2020\|EPI_ISL_513484\|2020 | ···················· | ···················· | ···················· | ···················· | ···················· | ···················· |
| F | 3770 | hCoV-19/USA/ME-HETL-J0074/2020\|EPI_ISL_513485\|2020 | ···················· | ···················· | ···················· | ···················· | ···················· | ···················· |
| F | 3771 | hCoV-19/USA/ME-HETL-J0075/2020\|EPI_ISL_513486\|2020 | ···················· | ···················· | ···················· | ···················· | ···················· | ···················· |
| F | 3772 | hCoV-19/USA/ME-HETL-J0077/2020\|EPI_ISL_513488\|2020 | ···················· | ···················· | ···················· | ···················· | ···················· | ···················· |
| F | 3773 | hCoV-19/USA/ME-HETL-J0078/2020\|EPI_ISL_513489\|2020 | ···················· | ···················· | ···················· | ···················· | ···················· | ···················· |
| F | 3774 | hCoV-19/USA/ME-HETL-J0079/2020\|EPI_ISL_513490\|2020 | ···················· | ···················· | ···················· | ···················· | ···················· | ···················· |
| F | 3775 | hCoV-19/USA/ME-HETL-J0080/2020\|EPI_ISL_513491\|2020 | ···················· | ···················· | ···················· | ···················· | ···················· | ···················· |
| F | 3776 | hCoV-19/USA/ME-HETL-J0087/2020\|EPI_ISL_513496\|2020 | ···················· | ···················· | ···················· | ···················· | ···················· | ···················· |
| F | 3777 | hCoV-19/USA/ME-HETL-J0088/2020\|EPI_ISL_513497\|2020 | ···················· | ···················· | ···················· | ···················· | ···················· | ···················· |
| F | 3778 | hCoV-19/USA/ME-HETL-J0089/2020\|EPI_ISL_513498\|2020 | ···················· | ···················· | ···················· | ···················· | ···················· | ···················· |
| F | 3779 | hCoV-19/USA/ME-HETL-J0090/2020\|EPI_ISL_513499\|2020 | ···················· | ···················· | ···················· | ···················· | ···················· | ···················· |
| F | 3780 | hCoV-19/India/GJ-GBRC353/2020\|EPI_ISL_514600\|2020- | ···················· | ···················· | ···················· | ···················· | ···················· | ···················· |
| F | 3781 | hCoV-19/India/GJ-GBRC354a/2020\|EPI_ISL_514601\|2020 | ···················· | ···················· | ···················· | ···················· | ···················· | ···················· |
| F | 3782 | hCoV-19/India/GJ-GBRC354b/2020\|EPI_ISL_514602\|2020 | ···················· | ···················· | ···················· | ···················· | ···················· | ···················· |
| F | 3783 | hCoV-19/India/GJ-GBRC355a/2020\|EPI_ISL_514603\|2020 | ···················· | ·A·················· | ···················· | ···················· | ···················· | ··A················· |
| F | 3784 | hCoV-19/USA/MN-MDH-1418/2020\|EPI_ISL_514637\|2020-0 | ···················· | ···················· | ···················· | ···················· | ···················· | ···················· |
| F | 3785 | hCoV-19/USA/MN-MDH-1419/2020\|EPI_ISL_514638\|2020-0 | ···················· | ···················· | ···················· | ···················· | ···················· | ···················· |
| F | 3786 | hCoV-19/USA/MN-MDH-1420/2020\|EPI_ISL_514639\|2020-0 | ···················· | ···················· | ···················· | ···················· | ···················· | ···················· |
| F | 3787 | hCoV-19/USA/MN-MDH-1425/2020\|EPI_ISL_514644\|2020-0 | ···················· | ···················· | ···················· | ···················· | ···················· | ···················· |
| F | 3788 | hCoV-19/USA/MN-MDH-1427/2020\|EPI_ISL_514646\|2020-0 | ···················· | ···················· | ···················· | ···················· | ···················· | ···················· |
| F | 3789 | hCoV-19/USA/MN-MDH-1428/2020\|EPI_ISL_514647\|2020-0 | ···················· | ···················· | ···················· | ···················· | ···················· | ···················· |
| F | 3790 | hCoV-19/USA/MN-MDH-1429/2020\|EPI_ISL_514648\|2020-0 | ···················· | ···················· | ···················· | ···················· | ···················· | ···················· |
| F | 3791 | hCoV-19/USA/MN-MDH-1430/2020\|EPI_ISL_514649\|2020-0 | ···················· | ···················· | ···················· | ···················· | ···················· | ···················· |
| F | 3792 | hCoV-19/USA/MN-MDH-1431/2020\|EPI_ISL_514650\|2020-0 | ···················· | ···················· | ···················· | ···················· | ···················· | ···················· |
| F | 3793 | hCoV-19/USA/MN-MDH-1432/2020\|EPI_ISL_514651\|2020-0 | ···················· | ···················· | ···················· | ···················· | ···················· | ···················· |
| F | 3794 | hCoV-19/USA/MN-MDH-1433/2020\|EPI_ISL_514652\|2020-0 | ···················· | ···················· | ···················· | ···················· | ···················· | ···················· |
| F | 3795 | hCoV-19/USA/MN-MDH-1434/2020\|EPI_ISL_514653\|2020-0 | ···················· | ···················· | ···················· | ···················· | ······T············· | ···················· |
| F | 3796 | hCoV-19/USA/MN-MDH-1435/2020\|EPI_ISL_514654\|2020-0 | ···················· | ···················· | ···················· | ···················· | ···················· | ···················· |
| F | 3797 | hCoV-19/USA/MN-MDH-1436/2020\|EPI_ISL_514655\|2020-0 | ···················· | ···················· | ···················· | ···················· | ···················· | ···················· |
| F | 3798 | hCoV-19/USA/MN-MDH-1437/2020\|EPI_ISL_514656\|2020-0 | ···················· | ···················· | ···················· | ···················· | ···················· | ···················· |
| F | 3799 | hCoV-19/USA/MN-MDH-1438/2020\|EPI_ISL_514657\|2020-0 | ···················· | ···················· | ···················· | ···················· | ···················· | ···················· |
| F | 3800 | hCoV-19/USA/MN-MDH-1439/2020\|EPI_ISL_514658\|2020-0 | ···················· | ···················· | ···················· | ···················· | ···················· | ···················· |
| F | 3801 | hCoV-19/USA/MN-MDH-1440/2020\|EPI_ISL_514659\|2020-0 | ···················· | ···················· | ···················· | ···················· | ···················· | ···················· |
| F | 3802 | hCoV-19/USA/MN-MDH-1441/2020\|EPI_ISL_514660\|2020-0 | ···················· | ···················· | ···················· | ···················· | ···················· | ···················· |
| F | 3803 | hCoV-19/USA/MN-MDH-1442/2020\|EPI_ISL_514661\|2020-0 | ···················· | ···················· | ···················· | ···················· | ···················· | ·····M·············· |
| F | 3804 | hCoV-19/USA/MN-MDH-1443/2020\|EPI_ISL_514662\|2020-0 | ···················· | ···················· | ···················· | ···················· | ···················· | ···················· |
| F | 3805 | hCoV-19/USA/MN-MDH-1444/2020\|EPI_ISL_514663\|2020-0 | ···················· | ···················· | ···················· | ···················· | ···················· | ···················· |
| F | 3806 | hCoV-19/USA/MN-MDH-1445/2020\|EPI_ISL_514664\|2020-0 | ···················· | ····G··············· | ···················· | ···················· | ···················· | ···················· |
| F | 3807 | hCoV-19/USA/MN-MDH-1446/2020\|EPI_ISL_514665\|2020-0 | ···················· | ···················· | ···················· | ···················· | ···················· | ···················· |
| F | 3808 | hCoV-19/USA/MN-MDH-1447/2020\|EPI_ISL_514666\|2020-0 | ···················· | ···················· | ···················· | ···················· | ···················· | ···················· |
| F | 3809 | hCoV-19/USA/MN-MDH-1448/2020\|EPI_ISL_514667\|2020-0 | ···················· | ····G··············· | ···················· | ···················· | ···················· | ···················· |
| F | 3810 | hCoV-19/USA/MN-MDH-1449/2020\|EPI_ISL_514668\|2020-0 | ···················· | ···················· | ···················· | ···················· | ···················· | ···················· |
| F | 3811 | hCoV-19/USA/MN-MDH-1450/2020\|EPI_ISL_514669\|2020-0 | ···················· | ···················· | ···················· | ···················· | ···················· | ···················· |
| F | 3812 | hCoV-19/USA/MN-MDH-1451/2020\|EPI_ISL_514670\|2020-0 | ···················· | ···················· | ···················· | ···················· | ···················· | ···················· |
| F | 3813 | hCoV-19/USA/MN-MDH-1452/2020\|EPI_ISL_514671\|2020-0 | ···················· | ···················· | ···················· | ···················· | ···················· | ···················· |
| F | 3814 | hCoV-19/USA/MN-MDH-1453/2020\|EPI_ISL_514672\|2020-0 | ···················· | ···················· | ···················· | ···················· | ···················· | ···················· |
| F | 3815 | hCoV-19/USA/NV-NSPHL-A0110/2020\|EPI_ISL_514673\|202 | ···················· | ···················· | ···················· | ···················· | ···················· | ···················· |
| F | 3816 | hCoV-19/USA/CA-CDPH007/2020\|EPI_ISL_515900\|2020-03 | ···················· | ···················· | ···················· | ···················· | ···················· | ···················· |
| F | 3817 | hCoV-19/USA/CA-CDPH008/2020\|EPI_ISL_515901\|2020-03 | ···················· | ···················· | ···················· | ···················· | ···················· | ···················· |
| F | 3818 | hCoV-19/USA/CA-CDPH009/2020\|EPI_ISL_515902\|2020-03 | ···················· | ···················· | ···················· | ···················· | ···················· | ···················· |
| F | 3819 | hCoV-19/USA/CA-CDPH010/2020\|EPI_ISL_515903\|2020-03 | ···················· | ···················· | ···················· | ···················· | ···················· | ···················· |
| F | 3820 | hCoV-19/USA/CA-CDPH012/2020\|EPI_ISL_515905\|2020-03 | ···················· | ···················· | ···················· | ···················· | ···················· | ···················· |
| F | 3821 | hCoV-19/USA/CA-CDPH013/2020\|EPI_ISL_515906\|2020-03 | ···················· | ···················· | ···················· | ···················· | ···················· | ···················· |
| F | 3822 | hCoV-19/USA/CA-CDPH014/2020\|EPI_ISL_515907\|2020-03 | ···················· | ···················· | ···················· | ···················· | ···················· | ···················· |
| F | 3823 | hCoV-19/USA/CA-CDPH015/2020\|EPI_ISL_515908\|2020-04 | ···················· | ···················· | ···················· | ···················· | ···················· | ···················· |
| F | 3824 | hCoV-19/USA/CA-CDPH016/2020\|EPI_ISL_515909\|2020-02 | ···················· | ···················· | ···················· | ···················· | ···················· | ···················· |
| F | 3825 | hCoV-19/USA/CA-CDPH017/2020\|EPI_ISL_515910\|2020-02 | ···················· | ···················· | ···················· | ···················· | ···················· | ···················· |
| F | 3826 | hCoV-19/USA/CA-CDPH018/2020\|EPI_ISL_515911\|2020-02 | ···················· | ···················· | ···················· | ···················· | ···················· | ···················· |
| F | 3827 | hCoV-19/USA/CA-CDPH020/2020\|EPI_ISL_515913\|2020-03 | ···················· | ···················· | ···················· | ···················· | ···················· | ···················· |
| F | 3828 | hCoV-19/USA/CA-CDPH025/2020\|EPI_ISL_515917\|2020-05 | ···················· | ···················· | ···················· | ···················· | ···················· | ···················· |
| F | 3829 | hCoV-19/USA/CA-CDPH026/2020\|EPI_ISL_515918\|2020-05 | ···················· | ···················· | ···················· | ···················· | ········Y··········· | ···················· |
| F | 3830 | hCoV-19/USA/CA-CDPH027/2020\|EPI_ISL_515919\|2020-05 | ···················· | ·A·················· | ···················· | ···················· | ···················· | ···················· |
| F | 3831 | hCoV-19/USA/CA-CDPH028/2020\|EPI_ISL_515920\|2020-05 | ···················· | ···················· | ···················· | ···················· | ···················· | ···················· |
| F | 3832 | hCoV-19/USA/CA-CDPH031/2020\|EPI_ISL_515922\|2020-06 | ···················· | ···················· | ···················· | ···················· | ···················· | ···················· |
| F | 3833 | hCoV-19/USA/CA-CDPH032/2020\|EPI_ISL_515923\|2020-06 | ···················· | ·A·················· | ···················· | ···················· | ···················· | ···················· |
| F | 3834 | hCoV-19/USA/CA-CDPH033/2020\|EPI_ISL_515924\|2020-06 | ···················· | ·A·················· | ···················· | ···················· | ···················· | ···················· |
| F | 3835 | hCoV-19/USA/CA-CDPH034/2020\|EPI_ISL_515925\|2020-06 | ···················· | ···················· | ···················· | ···················· | ···················· | ···················· |
| F | 3836 | hCoV-19/USA/CA-CDPH037/2020\|EPI_ISL_515928\|2020-06 | ···················· | ·A·················· | ···················· | ···················· | ···················· | ···················· |

| | | | | | | | | |
|---|---|---|---|---|---|---|---|---|
| F | 3837 | hCoV-19/USA/CA-CDPH038/2020\|EPI_ISL_515929\|2020-06 | ···················· | ···················· | ···················· | ···················· | ···················· | ···················· |
| F | 3838 | hCoV-19/India/KA-nimh-19510/2020\|EPI_ISL_515968\|20 | ···················· | ···················· | ···················· | ···················· | ···················· | ···················· |
| F | 3839 | hCoV-19/India/KA-nimh-19669/2020\|EPI_ISL_515969\|20 | ···················· | ···················· | ···················· | ···················· | ···················· | ···················· |
| F | 3840 | hCoV-19/India/KA-nimh-19688/2020\|EPI_ISL_515970\|20 | ···················· | ···················· | ···················· | ···················· | ···················· | ···················· |
| F | 3841 | hCoV-19/USA/WA-S2272/2020\|EPI_ISL_497700\|2020-06-1 | ···················· | ···················· | ···················· | ···················· | ···················· | ···················· |
| F | 3842 | hCoV-19/USA/WA-S2273/2020\|EPI_ISL_497701\|2020-06-0 | ···················· | ···················· | ···················· | ···················· | ···················· | ···················· |
| F | 3843 | hCoV-19/USA/WA-S2274/2020\|EPI_ISL_497702\|2020-06-0 | ···················· | ···················· | ···················· | ···················· | ···················· | ···················· |
| F | 3844 | hCoV-19/USA/WA-S2275/2020\|EPI_ISL_497703\|2020-06-0 | ···················· | ···················· | ···················· | ···················· | ···················· | ···················· |
| F | 3845 | hCoV-19/USA/WA-S2276/2020\|EPI_ISL_497704\|2020-06-0 | ···················· | ···················· | ···················· | ···················· | ···················· | ···················· |
| F | 3846 | hCoV-19/USA/WA-S2278/2020\|EPI_ISL_497706\|2020-06-0 | ···················· | ···················· | ···················· | ···················· | ···················· | ···················· |
| F | 3847 | hCoV-19/USA/WA-S2279/2020\|EPI_ISL_497707\|2020-06-1 | ···················· | ···················· | ···················· | ···················· | ···················· | ···················· |
| F | 3848 | hCoV-19/USA/WA-S2280/2020\|EPI_ISL_497708\|2020-06-0 | ···················· | ···················· | ···················· | ···················· | ···················· | ···················· |
| F | 3849 | hCoV-19/USA/WA-S2282/2020\|EPI_ISL_497710\|2020-06-1 | ···················· | ···················· | ···················· | ···················· | ···················· | ···················· |
| F | 3850 | hCoV-19/USA/WA-S2283/2020\|EPI_ISL_497711\|2020-06-1 | ···················· | ···················· | ···················· | ···················· | ···················· | ···················· |
| F | 3851 | hCoV-19/USA/WA-S2284/2020\|EPI_ISL_497712\|2020-06-1 | ···················· | ···················· | ···················· | ···················· | ···················· | ···················· |
| F | 3852 | hCoV-19/USA/WA-S2285/2020\|EPI_ISL_497713\|2020-06-1 | ···················· | ···················· | ···················· | ···················· | ···················· | ···················· |
| F | 3853 | hCoV-19/USA/WA-S2286/2020\|EPI_ISL_497714\|2020-06-1 | ···················· | ···················· | ···················· | ···················· | ···················· | ···················· |
| F | 3854 | hCoV-19/USA/WA-S2287/2020\|EPI_ISL_497715\|2020-06-1 | ···················· | ···················· | ···················· | ···················· | ···················· | ···················· |
| F | 3855 | hCoV-19/USA/WA-S2288/2020\|EPI_ISL_497716\|2020-06-1 | ···················· | ···················· | ···················· | ···················· | ···················· | ···················· |
| F | 3856 | hCoV-19/USA/WA-S2289/2020\|EPI_ISL_497717\|2020-06-0 | ···················· | ···················· | ···················· | ···················· | ···················· | ···················· |
| F | 3857 | hCoV-19/USA/WA-S2290/2020\|EPI_ISL_497718\|2020-06-0 | ···················· | ···················· | ···················· | ···················· | ···················· | ···················· |
| F | 3858 | hCoV-19/USA/WA-S2291/2020\|EPI_ISL_497719\|2020-06-0 | ···················· | ···················· | ···················· | ···················· | ···················· | ···················· |
| F | 3859 | hCoV-19/USA/WA-S2292/2020\|EPI_ISL_497720\|2020-06-0 | ···················· | ···················· | ···················· | ···················· | ···················· | ···················· |
| F | 3860 | hCoV-19/USA/WA-S2293/2020\|EPI_ISL_497721\|2020-06-1 | ···················· | ···················· | ···················· | ···················· | ···················· | ···················· |
| F | 3861 | hCoV-19/USA/WA-S2294/2020\|EPI_ISL_497722\|2020-06-1 | ···················· | ···················· | ···················· | ···················· | ···················· | ···················· |
| F | 3862 | hCoV-19/USA/WA-S2295/2020\|EPI_ISL_497723\|2020-06-1 | ···················· | ···················· | ···················· | ···················· | ···················· | ···················· |
| F | 3863 | hCoV-19/USA/WA-S2296/2020\|EPI_ISL_497724\|2020-06-1 | ···················· | ···················· | ···················· | ···················· | ···················· | ···················· |
| F | 3864 | hCoV-19/USA/WA-S2297/2020\|EPI_ISL_497725\|2020-06-1 | ···················· | ···················· | ···················· | ···················· | ···················· | ···················· |
| F | 3865 | hCoV-19/USA/WA-S2298/2020\|EPI_ISL_497726\|2020-06-1 | ···················· | ···················· | ···················· | ···················· | ···················· | ···················· |
| F | 3866 | hCoV-19/Colombia/RIS-INS-81402/2020\|EPI_ISL_497736 | ···················· | ···················· | ···················· | ···················· | ···················· | ···················· |
| F | 3867 | hCoV-19/Colombia/BOL-INS-84730/2020\|EPI_ISL_497738 | ···················· | ···················· | ···················· | ···················· | ···················· | ···················· |
| F | 3868 | hCoV-19/Colombia/QUI-INS-85082/2020\|EPI_ISL_497744 | ···················· | ···················· | ···················· | ···················· | ···················· | ···················· |
| F | 3869 | hCoV-19/Colombia/RIS-INS-87267/2020\|EPI_ISL_497745 | ···················· | ···················· | ···················· | ···················· | ···················· | ···················· |
| F | 3870 | hCoV-19/India/UN-N5/2020\|EPI_ISL_497758\|2020 | ···················· | ···················· | ···················· | ···················· | ····N····N·········· | ···················· |
| F | 3871 | hCoV-19/India/UN-N6/2020\|EPI_ISL_497760\|2020 | ···················· | ···················· | ···················· | ···················· | ····N····N·········· | ···N············N··· |
| F | 3873 | hCoV-19/India/UN-N8/2020\|EPI_ISL_497763\|2020 | ·····N··N··········· | ·A·················· | ···················· | ···················· | ···················· | ···A················ |
| F | 3874 | hCoV-19/India/UN-N9/2020\|EPI_ISL_497764\|2020 | ···················· | ···················· | ···················· | ···················· | ····N····N·········· | ···N············N··· |
| F | 3875 | hCoV-19/India/UN-N10/2020\|EPI_ISL_497765\|2020 | ···················· | ·A·················· | ···················· | ···················· | ····N····N·········· | ···A················ |
| F | 3876 | hCoV-19/India/UN-A349/2020\|EPI_ISL_497766\|2020 | ···················· | ·A·················· | ···················· | ···················· | ····N····N·········· | ···A················ |
| F | 3877 | hCoV-19/India/UN-10270/2020\|EPI_ISL_497767\|2020 | ···················· | ···················· | ···················· | ···················· | ····N····N·········· | ·····N·············· |
| F | 3878 | hCoV-19/Hong Kong/HKU-200723-002/2020\|EPI_ISL_4977 | ···················· | ···················· | ···················· | ···················· | ···················· | ···················· |
| F | 3879 | hCoV-19/Hong Kong/HKU-200723-003/2020\|EPI_ISL_4977 | ···················· | ···················· | ···················· | ···················· | ···················· | ···················· |
| F | 3880 | hCoV-19/Hong Kong/HKU-200723-004/2020\|EPI_ISL_4977 | ···················· | ···················· | ···················· | ···················· | ···················· | ···················· |
| F | 3881 | hCoV-19/Hong Kong/HKU-200723-006/2020\|EPI_ISL_4977 | ···················· | ···················· | ···················· | ···················· | ···················· | ···················· |
| F | 3882 | hCoV-19/Hong Kong/HKU-200723-015/2020\|EPI_ISL_4977 | ···················· | ···················· | ···················· | ···················· | ···················· | ···················· |
| F | 3883 | hCoV-19/Hong Kong/HKU-200723-016/2020\|EPI_ISL_4977 | ···················· | ···················· | ···················· | ···················· | ···················· | ···················· |
| F | 3884 | hCoV-19/Hong Kong/HKU-200723-017/2020\|EPI_ISL_4977 | ···················· | ···················· | ···················· | ···················· | ···················· | ···················· |
| F | 3885 | hCoV-19/Hong Kong/HKU-200723-018/2020\|EPI_ISL_4977 | ···················· | ···················· | ···················· | ···················· | ···················· | ···················· |
| F | 3886 | hCoV-19/Hong Kong/HKU-200723-019/2020\|EPI_ISL_4977 | ···················· | ···················· | ···················· | ···················· | ···················· | ···················· |
| F | 3887 | hCoV-19/Hong Kong/HKU-200723-023/2020\|EPI_ISL_4977 | ···················· | ···················· | ···················· | ···················· | ···················· | ···················· |
| F | 3888 | hCoV-19/Hong Kong/HKU-200723-024/2020\|EPI_ISL_4977 | ···················· | ···················· | ···················· | ···················· | ···················· | ···················· |
| F | 3889 | hCoV-19/Hong Kong/HKU-200723-026/2020\|EPI_ISL_4977 | ···················· | ···················· | ···················· | ···················· | ···················· | ···················· |
| F | 3890 | hCoV-19/Hong Kong/HKU-200723-027/2020\|EPI_ISL_4977 | ···················· | ···················· | ···················· | ···················· | ···················· | ···················· |
| F | 3891 | hCoV-19/Hong Kong/HKU-200723-028/2020\|EPI_ISL_4977 | ···················· | ···················· | ···················· | ···················· | ···················· | ···················· |
| F | 3892 | hCoV-19/Hong Kong/HKU-200723-029/2020\|EPI_ISL_4977 | ···················· | ···················· | ···················· | ···················· | ···················· | ···················· |
| F | 3893 | hCoV-19/Hong Kong/HKU-200723-030/2020\|EPI_ISL_4977 | ···················· | ···················· | ···················· | ···················· | ···················· | ···················· |
| F | 3894 | hCoV-19/Hong Kong/HKU-200723-031/2020\|EPI_ISL_4977 | ···················· | ···················· | ···················· | ···················· | ···················· | ···················· |
| F | 3895 | hCoV-19/Hong Kong/HKU-200723-032/2020\|EPI_ISL_4977 | ···················· | ···················· | ···················· | ···················· | ···················· | ···················· |
| F | 3896 | hCoV-19/England/LIVE-A263B/2020\|EPI_ISL_500000\|202 | ···················· | ···················· | ···················· | ···················· | ···················· | ···················· |
| F | 3897 | hCoV-19/England/LIVE-A2659/2020\|EPI_ISL_500001\|202 | ···················· | ···················· | ···················· | ···················· | ···················· | ···················· |
| F | 3898 | hCoV-19/England/LIVE-A26D1/2020\|EPI_ISL_500002\|202 | ···················· | ···················· | ···················· | ···················· | ···················· | ···················· |
| F | 3899 | hCoV-19/England/LIVE-A26E0/2020\|EPI_ISL_500003\|202 | ···················· | ···················· | ···················· | ···················· | ···················· | ···················· |
| F | 3900 | hCoV-19/England/LIVE-A26FF/2020\|EPI_ISL_500004\|202 | ···················· | ···················· | ···················· | ···················· | ···················· | ···················· |
| F | 3901 | hCoV-19/England/LIVE-A270B/2020\|EPI_ISL_500005\|202 | ···················· | ···················· | ···················· | ···················· | ···················· | ···················· |
| F | 3902 | hCoV-19/England/LIVE-A2756/2020\|EPI_ISL_500006\|202 | ···················· | ···················· | ···················· | ···················· | ···················· | ···················· |
| F | 3908 | hCoV-19/England/LIVE-A4912/2020\|EPI_ISL_500055\|202 | ···················· | ···················· | ···················· | ···················· | ···················· | ···················· |
| F | 3910 | hCoV-19/England/LIVE-A496D/2020\|EPI_ISL_500057\|202 | ···················· | ···················· | ···················· | ···················· | ···················· | ···················· |
| F | 3911 | hCoV-19/England/LIVE-A498B/2020\|EPI_ISL_500058\|202 | ···················· | ···················· | ···················· | ···················· | ···················· | ···················· |
| F | 3917 | hCoV-19/England/LIVE-A4A6A/2020\|EPI_ISL_500064\|202 | ···················· | ···················· | ···················· | ···················· | ···················· | ···················· |
| F | 3920 | hCoV-19/England/LIVE-A4AE2/2020\|EPI_ISL_500071\|202 | ···················· | ···················· | ···················· | ···················· | ···················· | ···················· |
| F | 3921 | hCoV-19/England/LIVE-A4B0D/2020\|EPI_ISL_500072\|202 | ···················· | ···················· | ···················· | ···················· | ···················· | ···················· |
| F | 3922 | hCoV-19/England/LIVE-A4B2B/2020\|EPI_ISL_500073\|202 | ···················· | ···················· | ···················· | ···················· | ···················· | ···················· |
| F | 3932 | hCoV-19/England/LIVE-A4CDD/2020\|EPI_ISL_500083\|202 | ···················· | ···················· | ···················· | ···················· | ···················· | ···················· |
| F | 3933 | hCoV-19/England/LIVE-A4D16/2020\|EPI_ISL_500084\|202 | ···················· | ···················· | ···················· | ···················· | ···················· | ···················· |
| F | 3934 | hCoV-19/England/LIVE-A4D52/2020\|EPI_ISL_500085\|202 | ···················· | ···················· | ···················· | ···················· | ···················· | ···················· |
| F | 3935 | hCoV-19/England/LIVE-A4D70/2020\|EPI_ISL_500086\|202 | ···················· | ···················· | ···················· | ···················· | ···················· | ···················· |
| F | 3936 | hCoV-19/England/LIVE-A4D8F/2020\|EPI_ISL_500087\|202 | ···················· | ···················· | ···················· | ···················· | ···················· | ···················· |
| F | 3937 | hCoV-19/England/LIVE-A4DAD/2020\|EPI_ISL_500088\|202 | ···················· | ···················· | ···················· | ···················· | ···················· | ···················· |
| F | 3938 | hCoV-19/England/LIVE-A4DDA/2020\|EPI_ISL_500089\|202 | ···················· | ···················· | ···················· | ···················· | ···················· | ···················· |
| F | 3939 | hCoV-19/England/LIVE-A4DE9/2020\|EPI_ISL_500090\|202 | ···················· | ···················· | ···················· | ···················· | ···················· | ···················· |

| | | | | | | | | |
|---|---|---|---|---|---|---|---|---|
| F | 3940 | hCoV-19/England/LIVE-A4E04/2020\|EPI_ISL_500091\|202 | ···················· | ···················· | ···················· | ···················· | ···················· | ···················· |
| F | 3941 | hCoV-19/England/LIVE-A4E13/2020\|EPI_ISL_500092\|202 | ···················· | ···················· | ···················· | ···················· | ···················· | ···················· |
| F | 3942 | hCoV-19/England/LIVE-A4E7D/2020\|EPI_ISL_500093\|202 | ···················· | ···················· | ···················· | ···················· | ···················· | ···················· |
| F | 3945 | hCoV-19/England/LIVE-A4F01/2020\|EPI_ISL_500096\|202 | ···················· | ···················· | ···················· | ···················· | ···················· | ···················· |
| F | 3946 | hCoV-19/England/LIVE-A4F10/2020\|EPI_ISL_500097\|202 | ···················· | ···················· | ···················· | ···················· | ···················· | ···················· |
| F | 3948 | hCoV-19/England/LIVE-A4FB6/2020\|EPI_ISL_500099\|202 | ···················· | ···················· | ···················· | ···················· | ···················· | ···················· |
| F | 3949 | hCoV-19/Bangladesh/BCSIR-NILMRC-261/2020\|EPI_ISL_5 | ···················· | ···················· | ···················· | ···················· | ···················· | ···················· |
| F | 3950 | hCoV-19/USA/FL-BPHL-0596/2020\|EPI_ISL_508800\|2020- | ···················· | ···················· | ···················· | ···················· | ···················· | ···················· |
| F | 3951 | hCoV-19/USA/FL-BPHL-0597/2020\|EPI_ISL_508801\|2020- | ···················· | ···················· | ···················· | ···················· | ···················· | ···················· |
| F | 3952 | hCoV-19/USA/FL-BPHL-0598/2020\|EPI_ISL_508802\|2020- | ···················· | ···················· | ···················· | ···················· | ···················· | ···················· |
| F | 3953 | hCoV-19/USA/FL-BPHL-0599/2020\|EPI_ISL_508803\|2020- | ···················· | ···················· | ···················· | ···················· | ···················· | ···················· |
| F | 3954 | hCoV-19/USA/FL-BPHL-0600/2020\|EPI_ISL_508804\|2020- | ···················· | ···················· | ···················· | ···················· | ···················· | ···················· |
| F | 3955 | hCoV-19/USA/FL-BPHL-0601/2020\|EPI_ISL_508805\|2020- | ···················· | ···················· | ···················· | ···················· | ···················· | ···················· |
| F | 3956 | hCoV-19/USA/FL-BPHL-0603/2020\|EPI_ISL_508807\|2020- | ···················· | ···················· | ···················· | ···················· | ···················· | ···················· |
| F | 3957 | hCoV-19/USA/FL-BPHL-0617/2020\|EPI_ISL_508821\|2020- | ···················· | ···················· | ···················· | ···················· | ···················· | ···················· |
| F | 3959 | hCoV-19/USA/FL-BPHL-0633/2020\|EPI_ISL_508837\|2020- | ···················· | ···················· | ···················· | ···················· | ···················· | ···················· |
| F | 3960 | hCoV-19/USA/FL-BPHL-0634/2020\|EPI_ISL_508838\|2020- | ···················· | ···················· | ···················· | ···················· | ···················· | ···················· |
| F | 3961 | hCoV-19/USA/FL-BPHL-0635/2020\|EPI_ISL_508839\|2020- | ···················· | ···················· | ···················· | ···················· | ···················· | ···················· |
| F | 3962 | hCoV-19/USA/FL-BPHL-0636/2020\|EPI_ISL_508840\|2020- | ···················· | ···················· | ···················· | ···················· | ···················· | ···················· |
| F | 3963 | hCoV-19/USA/FL-BPHL-0637/2020\|EPI_ISL_508841\|2020- | ···················· | ···················· | ···················· | ···················· | ···················· | ···················· |
| F | 3964 | hCoV-19/Switzerland/ZH-ETHZ-220071/2020\|EPI_ISL_51 | ···················· | ···················· | ···················· | ···················· | ···················· | ···················· |
| F | 3965 | hCoV-19/Switzerland/GR-ETHZ-220073/2020\|EPI_ISL_51 | ···················· | ···················· | ···················· | ···················· | ···················· | ···················· |
| F | 3966 | hCoV-19/Switzerland/BS-ETHZ-220074/2020\|EPI_ISL_51 | ···················· | ···················· | ···················· | ···················· | ···················· | ···················· |
| F | 3967 | hCoV-19/Switzerland/ZH-ETHZ-220076/2020\|EPI_ISL_51 | ···················· | ···················· | ···················· | ···················· | ···················· | ···················· |
| F | 3968 | hCoV-19/Switzerland/GR-ETHZ-220077/2020\|EPI_ISL_51 | ···················· | ···················· | ···················· | ···················· | ···················· | ···················· |
| F | 3969 | hCoV-19/Switzerland/GR-ETHZ-220078/2020\|EPI_ISL_51 | ···················· | ···················· | ···················· | ···················· | ···················· | ···················· |
| F | 3970 | hCoV-19/Switzerland/BL-ETHZ-220079/2020\|EPI_ISL_51 | ···················· | ···················· | ···················· | ···················· | ···················· | ···················· |
| F | 3971 | hCoV-19/Switzerland/ZH-ETHZ-220080/2020\|EPI_ISL_51 | ···················· | ···················· | ···················· | ···················· | ···················· | ···················· |
| F | 3972 | hCoV-19/Switzerland/ZH-ETHZ-220081/2020\|EPI_ISL_51 | ···················· | ···················· | ···················· | ···················· | ···················· | ···················· |
| F | 3973 | hCoV-19/Switzerland/BE-ETHZ-220084/2020\|EPI_ISL_51 | ···················· | ·A·················· | ···················· | ···················· | ···················· | ···················· |
| F | 3974 | hCoV-19/Switzerland/BE-ETHZ-220085/2020\|EPI_ISL_51 | ···················· | ···················· | ···················· | ···················· | ···················· | ···················· |
| F | 3975 | hCoV-19/Switzerland/VD-ETHZ-220087/2020\|EPI_ISL_51 | ···················· | ···················· | ···················· | ···················· | ···················· | ···················· |
| F | 3976 | hCoV-19/Switzerland/BL-ETHZ-220088/2020\|EPI_ISL_51 | ···················· | ···················· | ···················· | ···················· | ···················· | ···················· |
| F | 3977 | hCoV-19/Switzerland/GR-ETHZ-220089/2020\|EPI_ISL_51 | ···················· | ···················· | ···················· | ···················· | ···················· | ···················· |
| F | 3978 | hCoV-19/India/GJ-GBRC324/2020\|EPI_ISL_512058\|2020- | ···················· | ·A·················· | ···················· | ···················· | ···················· | ···A················ |
| F | 3979 | hCoV-19/India/GJ-GBRC325/2020\|EPI_ISL_512059\|2020- | ···················· | ·A·················· | ···················· | ···················· | ···················· | ···A················ |
| F | 3980 | hCoV-19/India/GJ-GBRC326/2020\|EPI_ISL_512060\|2020- | ···················· | ·A·················· | ···················· | ···················· | ···················· | ···A················ |
| F | 3981 | hCoV-19/India/GJ-GBRC328/2020\|EPI_ISL_512063\|2020- | ···················· | ·A·················· | ···················· | ···················· | ···················· | ···A················ |
| F | 3982 | hCoV-19/India/GJ-GBRC329/2020\|EPI_ISL_512064\|2020- | ···················· | ·A·················· | ···················· | ···················· | ···················· | ···A················ |
| F | 3983 | hCoV-19/India/GJ-GBRC330/2020\|EPI_ISL_512065\|2020- | ···················· | ·A·················· | ···················· | ···················· | ···················· | ···A················ |
| F | 3984 | hCoV-19/India/GJ-GBRC331/2020\|EPI_ISL_512066\|2020- | ···T················ | ·A·················· | ···················· | ···················· | ···················· | ···A················ |
| F | 3985 | hCoV-19/India/GJ-GBRC332a/2020\|EPI_ISL_512067\|2020 | ···················· | ·A·················· | ···················· | ···················· | ···················· | ···A················ |
| F | 3986 | hCoV-19/India/GJ-GBRC332b/2020\|EPI_ISL_512068\|2020 | ···················· | ·A·················· | ···················· | ···················· | ···················· | ···A················ |
| F | 3987 | hCoV-19/India/GJ-GBRC333/2020\|EPI_ISL_512069\|2020- | ···················· | ·A·················· | ···················· | ···················· | ···················· | ···A················ |
| F | 3988 | hCoV-19/India/GJ-GBRC334a/2020\|EPI_ISL_512070\|2020 | ···················· | ···················· | ···················· | ···················· | ···················· | ···················· |
| F | 3989 | hCoV-19/India/GJ-GBRC334b/2020\|EPI_ISL_512071\|2020 | ···················· | ···················· | ···················· | ···················· | ···················· | ···················· |
| F | 3990 | hCoV-19/India/GJ-GBRC335a/2020\|EPI_ISL_512072\|2020 | ···················· | ·A·················· | ···················· | ···················· | ···················· | ···A················ |
| F | 3991 | hCoV-19/India/GJ-GBRC335b/2020\|EPI_ISL_512073\|2020 | ···················· | ·A·················· | ···················· | ···················· | ···················· | ···A················ |
| F | 3992 | hCoV-19/India/GJ-GBRC336/2020\|EPI_ISL_512074\|2020- | ···················· | ·A·················· | ···················· | ···················· | ···················· | ···A················ |
| F | 3993 | hCoV-19/India/GJ-GBRC338a/2020\|EPI_ISL_512075\|2020 | ···················· | ···················· | ···················· | ···················· | ···················· | ···················· |
| F | 3994 | hCoV-19/India/GJ-GBRC338b/2020\|EPI_ISL_512076\|2020 | ···················· | ···················· | ···················· | ···················· | ···················· | ···················· |
| F | 3995 | hCoV-19/India/GJ-GBRC-339/2020\|EPI_ISL_512077\|2020 | ···················· | ···················· | ···················· | ···················· | ···················· | ···················· |
| F | 3996 | hCoV-19/USA/WA-UW-10116/2020\|EPI_ISL_512086\|2020-0 | ···················· | ···················· | ···················· | ···················· | ···················· | ···················· |
| F | 3997 | hCoV-19/Ireland/KE-NVRL-71IRL67942/2020\|EPI_ISL_51 | ···················· | ···················· | ···················· | ···················· | ···················· | ···················· |
| F | 3998 | hCoV-19/Ireland/MH-NVRL-71IRL72075/2020\|EPI_ISL_51 | ···················· | ···················· | ···················· | ···················· | ···················· | ···················· |
| F | 3999 | hCoV-19/Ireland/D-NVRL-71IRL72362/2020\|EPI_ISL_512 | ···················· | ···················· | ···················· | ···················· | ···················· | ···················· |
| F | 4000 | hCoV-19/Ireland/KE-NVRL-71IRL74566/2020\|EPI_ISL_51 | ···················· | ···················· | ···················· | ···················· | ···················· | ···················· |
| F | 4001 | hCoV-19/Ireland/D-NVRL-71IRL74709/2020\|EPI_ISL_512 | ···················· | ···················· | ···················· | ···················· | ···················· | ···················· |
| F | 4002 | hCoV-19/Ireland/CO-NVRL-71IRL77106/2020\|EPI_ISL_51 | ···················· | ·A·················· | ···················· | ···················· | ···················· | ···A················ |
| F | 4003 | hCoV-19/Ireland/D-NVRL-71IRL79618/2020\|EPI_ISL_512 | ···················· | ···················· | ···················· | ···················· | ···················· | ···················· |
| F | 4004 | hCoV-19/Ireland/KE-NVRL-71IRL80058/2020\|EPI_ISL_51 | ···················· | ···················· | ···················· | ···················· | ···················· | ···················· |
| F | 4005 | hCoV-19/Ireland/LH-NVRL-71IRL81038/2020\|EPI_ISL_51 | ···················· | ···················· | ···················· | ···················· | ···················· | ···················· |
| F | 4006 | hCoV-19/Ireland/LS-NVRL-71IRL81151/2020\|EPI_ISL_51 | ···················· | ···················· | ···················· | ···················· | ···················· | ···················· |
| F | 4007 | hCoV-19/United Kingdom/USAFSAM-S026/2020\|EPI_ISL_5 | ···················· | ···················· | ···················· | ···················· | ···················· | ···················· |
| F | 4008 | hCoV-19/Cuba/USAFSAM-S030/2020\|EPI_ISL_513312\|2020 | ···················· | ···················· | ···················· | ···················· | ···················· | ···················· |
| F | 4009 | hCoV-19/Australia/NSW672/2020\|EPI_ISL_513315\|2020- | ···················· | ···················· | ···················· | ···················· | ···················· | ···················· |
| F | 4010 | hCoV-19/Australia/NSW673/2020\|EPI_ISL_513316\|2020- | ···················· | ···················· | ···················· | ···················· | ···················· | ···················· |
| F | 4011 | hCoV-19/Australia/NSW727/2020\|EPI_ISL_513370\|2020- | ···················· | ···················· | ···················· | ···················· | ···················· | ···················· |
| F | 4012 | hCoV-19/Australia/NSW728/2020\|EPI_ISL_513371\|2020- | ···················· | ···················· | ···················· | ···················· | ···················· | ···················· |
| F | 4013 | hCoV-19/Australia/NSW729/2020\|EPI_ISL_513372\|2020- | ···················· | ···················· | ···················· | ···················· | ···················· | ···················· |
| F | 4014 | hCoV-19/Australia/NSW730/2020\|EPI_ISL_513373\|2020- | ···················· | ···················· | ···················· | ···················· | ···················· | ···················· |
| F | 4015 | hCoV-19/Australia/NSW731/2020\|EPI_ISL_513374\|2020- | ···················· | ···················· | ···················· | ···················· | ···················· | ···················· |
| F | 4016 | hCoV-19/Australia/NSW732/2020\|EPI_ISL_513375\|2020- | ···················· | ···················· | ···················· | ···················· | ···················· | ···················· |
| F | 4017 | hCoV-19/Australia/NSW733/2020\|EPI_ISL_513376\|2020- | ···················· | ···················· | ···················· | ···················· | ···················· | ···················· |
| F | 4018 | hCoV-19/Australia/NSW734/2020\|EPI_ISL_513377\|2020- | ···················· | ···················· | ···················· | ···················· | ···················· | ···················· |
| F | 4019 | hCoV-19/Australia/NSW735/2020\|EPI_ISL_513378\|2020- | ···················· | ···················· | ···················· | ···················· | ···················· | ···················· |
| F | 4020 | hCoV-19/Australia/NSW736/2020\|EPI_ISL_513379\|2020- | ···················· | ···················· | ···················· | ···················· | ···················· | ···················· |
| F | 4021 | hCoV-19/Australia/NSW737/2020\|EPI_ISL_513380\|2020- | ···················· | ···················· | ···················· | ···················· | ···················· | ···················· |
| F | 4022 | hCoV-19/Australia/NSW738/2020\|EPI_ISL_513381\|2020- | ···················· | ···················· | ···················· | ···················· | ···················· | ···················· |
| F | 4023 | hCoV-19/Australia/NSW739/2020\|EPI_ISL_513382\|2020- | ···················· | ···················· | ···················· | ···················· | ···················· | ···················· |

| | | | | | | | |
|---|---|---|---|---|---|---|---|
| F | 4024 hCoV-19/Australia/NSW740/2020\|EPI_ISL_513383\|2020- | ···················· | ···················· | ···················· | ···················· | ···················· | ···················· |
| F | 4025 hCoV-19/Australia/NSW741/2020\|EPI_ISL_513384\|2020- | ···················· | ···················· | ···················· | ···················· | ···················· | ···················· |
| F | 4026 hCoV-19/Australia/NSW742/2020\|EPI_ISL_513385\|2020- | ···················· | ···················· | ···················· | ···················· | ···················· | ···················· |
| F | 4027 hCoV-19/Australia/NSW743/2020\|EPI_ISL_513386\|2020- | ···················· | ···················· | ···················· | ···················· | ···················· | ···················· |
| F | 4028 hCoV-19/Australia/NSW744/2020\|EPI_ISL_513387\|2020- | ···················· | ···················· | ···················· | ···················· | ···················· | ···················· |
| F | 4029 hCoV-19/Australia/NSW745/2020\|EPI_ISL_513388\|2020- | ···················· | ···················· | ···················· | ···················· | ···················· | ···················· |
| F | 4030 hCoV-19/Australia/NSW747/2020\|EPI_ISL_513389\|2020- | ···················· | ···················· | ···················· | ···················· | ···················· | ···················· |
| F | 4031 hCoV-19/Australia/NSW748/2020\|EPI_ISL_513390\|2020- | ···················· | ···················· | ···················· | ···················· | ···················· | ···················· |
| F | 4032 hCoV-19/Australia/NSW749/2020\|EPI_ISL_513391\|2020- | ···················· | ···················· | ···················· | ···················· | ···················· | ···················· |
| F | 4033 hCoV-19/Australia/NSW750/2020\|EPI_ISL_513392\|2020- | ···················· | ···················· | ···················· | ···················· | ···················· | ···················· |
| F | 4034 hCoV-19/Australia/NSW751/2020\|EPI_ISL_513393\|2020- | ···················· | ···················· | ···················· | ···················· | ···················· | ···················· |
| F | 4035 hCoV-19/Australia/NSW752/2020\|EPI_ISL_513394\|2020- | ···················· | ···················· | ···················· | ···················· | ···················· | ···················· |
| F | 4036 hCoV-19/Australia/NSW753/2020\|EPI_ISL_513395\|2020- | ···················· | ···················· | ···················· | ···················· | ···················· | ···················· |
| F | 4037 hCoV-19/Australia/NSW754/2020\|EPI_ISL_513396\|2020- | ···················· | ···················· | ···················· | ···················· | ···················· | ···················· |
| F | 4038 hCoV-19/Australia/NSW755/2020\|EPI_ISL_513397\|2020- | ···················· | ···················· | ···················· | ···················· | ···················· | ···················· |
| F | 4039 hCoV-19/Australia/NSW756/2020\|EPI_ISL_513398\|2020- | ···················· | ···················· | ···················· | ···················· | ···················· | ···················· |
| F | 4040 hCoV-19/Australia/NSW757/2020\|EPI_ISL_513399\|2020- | ···················· | ···················· | ···················· | ···················· | ···················· | ···················· |
| F | 4041 hCoV-19/India/GJ-GBRC355b/2020\|EPI_ISL_514604\|2020 | ···················· | ·A·················· | ···················· | ···················· | ···················· | ··A················· |
| F | 4042 hCoV-19/India/GJ-GBRC356/2020\|EPI_ISL_514605\|2020- | ···················· | ·A·················· | ···················· | ···················· | ···················· | ··A················· |
| F | 4043 hCoV-19/India/GJ-GBRC357a/2020\|EPI_ISL_514606\|2020 | ···················· | ·A·················· | ···················· | ···················· | ···················· | ··A················· |
| F | 4044 hCoV-19/India/GJ-GBRC357b/2020\|EPI_ISL_514607\|2020 | ···················· | ·A·················· | ···················· | ···················· | ···················· | ··A················· |
| F | 4045 hCoV-19/India/GJ-GBRC358/2020\|EPI_ISL_514608\|2020- | ···················· | ·A·················· | ···················· | ···················· | ···················· | ··A················· |
| F | 4046 hCoV-19/India/GJ-GBRC359/2020\|EPI_ISL_514609\|2020- | ···················· | ·A·················· | ···················· | ···················· | ···················· | ··A················· |
| F | 4047 hCoV-19/India/GJ-GBRC360/2020\|EPI_ISL_514610\|2020- | ···················· | ···················· | ···················· | ···················· | ···················· | ···················· |
| F | 4048 hCoV-19/India/GJ-GBRC361a/2020\|EPI_ISL_514611\|2020 | ···················· | ·A·················· | ···················· | ···················· | ···················· | ··A················· |
| F | 4049 hCoV-19/India/GJ-GBRC361b/2020\|EPI_ISL_514612\|2020 | ···················· | ·A·················· | ···················· | ···················· | ···················· | ··A················· |
| F | 4050 hCoV-19/Bangladesh/NGRI-NSTU-18/2020\|EPI_ISL_51461 | ···················· | ···················· | ···················· | ···················· | ···················· | ···················· |
| F | 4051 hCoV-19/Bangladesh/NGRI-NSTU-21/2020\|EPI_ISL_51461 | ···················· | ·A·················· | ···················· | ···················· | ···················· | ··A················· |
| F | 4052 hCoV-19/USA/MN-MDH-1397/2020\|EPI_ISL_514616\|2020-0 | ···················· | ···················· | ···················· | ···················· | ···················· | ···················· |
| F | 4053 hCoV-19/USA/MN-MDH-1398/2020\|EPI_ISL_514617\|2020-0 | ···················· | ···················· | ···················· | ···················· | ···················· | ···················· |
| F | 4054 hCoV-19/USA/MN-MDH-1399/2020\|EPI_ISL_514618\|2020-0 | ···················· | ···················· | ···················· | ···················· | ···················· | ···················· |
| F | 4055 hCoV-19/USA/MN-MDH-1400/2020\|EPI_ISL_514619\|2020-0 | ···················· | ···················· | ···················· | ···················· | ···················· | ···················· |
| F | 4056 hCoV-19/USA/MN-MDH-1401/2020\|EPI_ISL_514620\|2020-0 | ···················· | ···················· | ···················· | ···················· | ···················· | ···················· |
| F | 4057 hCoV-19/USA/MN-MDH-1402/2020\|EPI_ISL_514621\|2020-0 | ···················· | ···················· | ···················· | ···················· | ···················· | ···················· |
| F | 4058 hCoV-19/USA/MN-MDH-1403/2020\|EPI_ISL_514622\|2020-0 | ···················· | ···················· | ···················· | ···················· | ···················· | ···················· |
| F | 4059 hCoV-19/USA/MN-MDH-1404/2020\|EPI_ISL_514623\|2020-0 | ···················· | ···················· | ···················· | ···················· | ··N················· | ···················· |
| F | 4060 hCoV-19/USA/MN-MDH-1405/2020\|EPI_ISL_514624\|2020-0 | ···················· | ···················· | ···················· | ···················· | ···················· | ···················· |
| F | 4061 hCoV-19/USA/MN-MDH-1406/2020\|EPI_ISL_514625\|2020-0 | ···················· | ···················· | ···················· | ···················· | ···················· | ···················· |
| F | 4062 hCoV-19/USA/MN-MDH-1407/2020\|EPI_ISL_514626\|2020-0 | ···················· | ···················· | ···················· | ···················· | ···················· | ···················· |
| F | 4063 hCoV-19/USA/MN-MDH-1408/2020\|EPI_ISL_514627\|2020-0 | ···················· | ·A·················· | ···················· | ···················· | ···················· | ···················· |
| F | 4064 hCoV-19/USA/MN-MDH-1409/2020\|EPI_ISL_514628\|2020-0 | ···················· | ···················· | ···················· | ···················· | ···················· | ···················· |
| F | 4065 hCoV-19/USA/MN-MDH-1410/2020\|EPI_ISL_514629\|2020-0 | ···················· | ···················· | ···················· | ···················· | ···················· | ···················· |
| F | 4066 hCoV-19/USA/MN-MDH-1411/2020\|EPI_ISL_514630\|2020-0 | ···················· | ···················· | ···················· | ···················· | ···················· | ···················· |
| F | 4067 hCoV-19/USA/MN-MDH-1412/2020\|EPI_ISL_514631\|2020-0 | ···················· | ···················· | ···················· | ···················· | ···················· | ···················· |
| F | 4068 hCoV-19/USA/MN-MDH-1413/2020\|EPI_ISL_514632\|2020-0 | ···················· | ···················· | ···················· | ···················· | ···················· | ···················· |
| F | 4069 hCoV-19/USA/MN-MDH-1414/2020\|EPI_ISL_514633\|2020-0 | ···················· | ···················· | ···················· | ···················· | ···················· | ···················· |
| F | 4070 hCoV-19/USA/MN-MDH-1415/2020\|EPI_ISL_514634\|2020-0 | ···················· | ···················· | ···················· | ···················· | ···················· | ···················· |
| F | 4071 hCoV-19/USA/MN-MDH-1416/2020\|EPI_ISL_514635\|2020-0 | ···················· | ·A·················· | ···················· | ···················· | ···················· | ···················· |
| F | 4072 hCoV-19/USA/MN-MDH-1417/2020\|EPI_ISL_514636\|2020-0 | ···················· | ···················· | ···················· | ···················· | ···················· | ···················· |
| F | 4073 hCoV-19/USA/MN-MDH-1421/2020\|EPI_ISL_514640\|2020-0 | ···················· | ···················· | ···················· | ···················· | ···················· | ···················· |
| F | 4074 hCoV-19/USA/MN-MDH-1422/2020\|EPI_ISL_514641\|2020-0 | ···················· | ···················· | ···················· | ···················· | ···················· | ···················· |
| F | 4075 hCoV-19/USA/MN-MDH-1423/2020\|EPI_ISL_514642\|2020-0 | ···················· | ···················· | ···················· | ···················· | ···················· | ···················· |
| F | 4076 hCoV-19/USA/MN-MDH-1424/2020\|EPI_ISL_514643\|2020-0 | ···················· | ···················· | ···················· | ···················· | ···················· | ···················· |
| F | 4077 hCoV-19/USA/MN-MDH-1426/2020\|EPI_ISL_514645\|2020-0 | ···················· | ···················· | ···················· | ···················· | ···················· | ···················· |
| F | 4078 hCoV-19/India/KA-nimh-3033/2020\|EPI_ISL_515938\|202 | ···················· | ···················· | ···················· | ···················· | ···················· | ···················· |
| F | 4079 hCoV-19/India/KA-nimh-3948/2020\|EPI_ISL_515941\|202 | ···················· | ···················· | ···················· | ···················· | ···················· | ···················· |
| F | 4080 hCoV-19/India/KA-nimh-3964/2020\|EPI_ISL_515942\|202 | ···················· | ···················· | ···················· | ···················· | ···················· | ···················· |
| F | 4081 hCoV-19/India/KA-nimh-11068/2020\|EPI_ISL_515945\|20 | ···················· | ···················· | ···················· | ···················· | ···················· | ···················· |
| F | 4082 hCoV-19/India/KA-nimh-11071/2020\|EPI_ISL_515947\|20 | ···················· | ···················· | ···················· | ···················· | ···················· | ···················· |
| F | 4083 hCoV-19/India/KA-nimh-11872/2020\|EPI_ISL_515952\|20 | ···················· | ···················· | ···················· | ···················· | ···················· | ···················· |
| F | 4084 hCoV-19/India/KA-nimh-13855/2020\|EPI_ISL_515954\|20 | ···················· | ···················· | ···················· | ···················· | ···················· | ···················· |
| F | 4085 hCoV-19/India/KA-nimh-14722/2020\|EPI_ISL_515957\|20 | ···················· | ···················· | ···················· | ···················· | ···················· | ···················· |
| F | 4086 hCoV-19/India/KA-nimh-14834/2020\|EPI_ISL_515958\|20 | ···················· | ···················· | ···················· | ···················· | ···················· | ···················· |
| F | 4087 hCoV-19/India/KA-nimh-14835/2020\|EPI_ISL_515959\|20 | ···················· | ···················· | ···················· | ···················· | ···················· | ···················· |
| F | 4088 hCoV-19/India/KA-nimh-15833/2020\|EPI_ISL_515961\|20 | ···················· | ···················· | ···················· | ···················· | ···················· | ···················· |
| F | 4089 hCoV-19/India/KA-nimh-17585/2020\|EPI_ISL_515963\|20 | ···················· | ···················· | ···················· | ···················· | ···················· | ···················· |
| F | 4090 hCoV-19/India/KA-nimh-17608/2020\|EPI_ISL_515964\|20 | ···················· | ···················· | ···················· | ···················· | ···················· | ···················· |
| F | 4091 hCoV-19/India/KA-nimh-19184/2020\|EPI_ISL_515966\|20 | ···················· | ···················· | ···················· | ···················· | ···················· | ···················· |
| F | 4092 hCoV-19/India/KA-nimh-19188/2020\|EPI_ISL_515967\|20 | ···················· | ···················· | ···················· | ···················· | ···················· | ···················· |
| F | 4093 hCoV-19/India/KA-nimh-19696/2020\|EPI_ISL_515971\|20 | ···················· | ···················· | ···················· | ···················· | ···················· | ···················· |
| F | 4094 hCoV-19/India/KA-nimh-19712/2020\|EPI_ISL_515972\|20 | ···················· | ···················· | ···················· | ···················· | ···················· | ···················· |
| F | 4095 hCoV-19/India/KA-nimh-19716/2020\|EPI_ISL_515973\|20 | ···················· | ···················· | ···················· | ···················· | ···················· | ···················· |
| F | 4096 hCoV-19/Denmark/ALAB-1000/2020\|EPI_ISL_515974\|2020 | ···················· | ···················· | ···················· | ···················· | ···················· | ···················· |
| F | 4097 hCoV-19/USA/WA-S1687/2020\|EPI_ISL_496113\|2020-05-3 | ···················· | ···················· | ···················· | ···················· | ···················· | ···················· |
| F | 4098 hCoV-19/USA/WA-S1688/2020\|EPI_ISL_496114\|2020-05-3 | ···················· | ···················· | ···················· | ···················· | ···················· | ···················· |
| F | 4099 hCoV-19/USA/WA-S1689/2020\|EPI_ISL_496115\|2020-05-3 | ···················· | ···················· | ···················· | ···················· | ···················· | ···················· |
| F | 4100 hCoV-19/USA/WA-S1690/2020\|EPI_ISL_496116\|2020-05-3 | ···················· | ···················· | ···················· | ···················· | ···················· | ···················· |
| F | 4101 hCoV-19/USA/WA-S1691/2020\|EPI_ISL_496117\|2020-05-3 | ···················· | ···················· | ···················· | ···················· | ···················· | ···················· |
| F | 4102 hCoV-19/USA/WA-S1692/2020\|EPI_ISL_496118\|2020-05-3 | ···················· | ···················· | ···················· | ···················· | ···················· | ···················· |
| F | 4103 hCoV-19/USA/WA-S1693/2020\|EPI_ISL_496119\|2020-05-3 | ···················· | ···················· | ···················· | ···················· | ···················· | ···················· |

| | | | | | | | | |
|---|---|---|---|---|---|---|---|---|
| F | 4104 | hCoV-19/USA/WA-S1694/2020\|EPI_ISL_496120\|2020-05-3 | ···················· | ···················· | ···················· | ···················· | ···················· | ···················· |
| F | 4105 | hCoV-19/USA/WA-S1696/2020\|EPI_ISL_496122\|2020-05-3 | ···················· | ···················· | ···················· | ···················· | ···················· | ···················· |
| F | 4106 | hCoV-19/USA/WA-S1697/2020\|EPI_ISL_496123\|2020-05-3 | ···················· | ···················· | ···················· | ···················· | ···················· | ···················· |
| F | 4107 | hCoV-19/USA/WA-S1698/2020\|EPI_ISL_496124\|2020-05-3 | ···················· | ···················· | ···················· | ···················· | ···················· | ···················· |
| F | 4108 | hCoV-19/USA/WA-S1699/2020\|EPI_ISL_496125\|2020-05-3 | ···················· | ···················· | ···················· | ···················· | ···················· | ···················· |
| F | 4109 | hCoV-19/USA/WA-S1700/2020\|EPI_ISL_496126\|2020-05-3 | ···················· | ···················· | ···················· | ···················· | ···················· | ···················· |
| F | 4110 | hCoV-19/USA/WA-S1701/2020\|EPI_ISL_496127\|2020-05-3 | ···················· | ···················· | ···················· | ···················· | ···················· | ···················· |
| F | 4111 | hCoV-19/USA/WA-S1702/2020\|EPI_ISL_496128\|2020-05-3 | ···················· | ···················· | ···················· | ···················· | ···················· | ···················· |
| F | 4112 | hCoV-19/USA/WA-S1703/2020\|EPI_ISL_496129\|2020-05-3 | ···················· | ···················· | ···················· | ···················· | ···················· | ···················· |
| F | 4113 | hCoV-19/USA/WA-S1704/2020\|EPI_ISL_496130\|2020-06-0 | ···················· | ···················· | ···················· | ···················· | ···················· | ···················· |
| F | 4114 | hCoV-19/USA/WA-S1706/2020\|EPI_ISL_496132\|2020-06-0 | ···················· | ···················· | ···················· | ···················· | ···················· | ···················· |
| F | 4115 | hCoV-19/USA/WA-S1707/2020\|EPI_ISL_496133\|2020-06-0 | ···················· | ···················· | ···················· | ···················· | ···················· | ···················· |
| F | 4116 | hCoV-19/USA/WA-S1708/2020\|EPI_ISL_496134\|2020-05-3 | ···················· | ···················· | ···················· | ···················· | ···················· | ···················· |
| F | 4117 | hCoV-19/USA/WA-S1731/2020\|EPI_ISL_496157\|2020-06-0 | ···················· | ···················· | ···················· | ···················· | ···················· | ···················· |
| F | 4118 | hCoV-19/USA/WA-S1732/2020\|EPI_ISL_496158\|2020-06-0 | ···················· | ···················· | ···················· | ···················· | ···················· | ···················· |
| F | 4119 | hCoV-19/USA/WA-S1733/2020\|EPI_ISL_496159\|2020-06-0 | ···················· | ···················· | ···················· | ···················· | ···················· | ···················· |
| F | 4120 | hCoV-19/USA/WA-S1734/2020\|EPI_ISL_496160\|2020-06-0 | ···················· | ···················· | ···················· | ···················· | ···················· | ···················· |
| F | 4121 | hCoV-19/USA/WA-S1735/2020\|EPI_ISL_496161\|2020-06-0 | ···················· | ···················· | ···················· | ···················· | ···················· | ···················· |
| F | 4122 | hCoV-19/USA/WA-S1736/2020\|EPI_ISL_496162\|2020-06-0 | ···················· | ···················· | ···················· | ···················· | ···················· | ···················· |
| F | 4123 | hCoV-19/USA/WA-S1737/2020\|EPI_ISL_496163\|2020-06-0 | ···················· | ···················· | ···················· | ···················· | ···················· | ···················· |
| F | 4124 | hCoV-19/USA/WA-S1738/2020\|EPI_ISL_496164\|2020-06-0 | ···················· | ···················· | ···················· | ···················· | ···················· | ···················· |
| F | 4125 | hCoV-19/USA/WA-S1739/2020\|EPI_ISL_496165\|2020-06-0 | ···················· | ···················· | ···················· | ···················· | ···················· | ···················· |
| F | 4126 | hCoV-19/USA/WA-S1740/2020\|EPI_ISL_496166\|2020-06-0 | ···················· | ···················· | ···················· | ···················· | ···················· | ···················· |
| F | 4127 | hCoV-19/USA/WA-S1741/2020\|EPI_ISL_496167\|2020-06-0 | ···················· | ···················· | ···················· | ···················· | ···················· | ···················· |
| F | 4128 | hCoV-19/USA/WA-S1742/2020\|EPI_ISL_496168\|2020-06-0 | ···················· | ···················· | ···················· | ···················· | ···················· | ···················· |
| F | 4129 | hCoV-19/USA/WA-S1743/2020\|EPI_ISL_496169\|2020-06-0 | ···················· | ···················· | ···················· | ···················· | ···················· | ···················· |
| F | 4130 | hCoV-19/USA/WA-S1745/2020\|EPI_ISL_496171\|2020-06-0 | ···················· | ···················· | ···················· | ···················· | ···················· | ···················· |
| F | 4131 | hCoV-19/USA/WA-S1747/2020\|EPI_ISL_496173\|2020-06-0 | ···················· | ···················· | ···················· | ···················· | ···················· | ···················· |
| F | 4132 | hCoV-19/USA/WA-S1748/2020\|EPI_ISL_496174\|2020-06-0 | ···················· | ···················· | ···················· | ···················· | ···················· | ···················· |
| F | 4133 | hCoV-19/USA/WA-S1749/2020\|EPI_ISL_496175\|2020-06-0 | ···················· | ···················· | ···················· | ···················· | ···················· | ···················· |
| F | 4134 | hCoV-19/USA/WA-S1750/2020\|EPI_ISL_496176\|2020-06-0 | ···················· | ···················· | ···················· | ···················· | ···················· | ···················· |
| F | 4135 | hCoV-19/USA/WA-S1751/2020\|EPI_ISL_496177\|2020-06-0 | ···················· | ···················· | ···················· | ···················· | ···················· | ···················· |
| F | 4136 | hCoV-19/USA/WA-S1752/2020\|EPI_ISL_496178\|2020-06-0 | ···················· | ···················· | ···················· | ···················· | ···················· | ···················· |
| F | 4137 | hCoV-19/USA/WA-S1753/2020\|EPI_ISL_496179\|2020-06-0 | ···················· | ···················· | ···················· | ···················· | ···················· | ···················· |
| F | 4138 | hCoV-19/USA/WA-S1754/2020\|EPI_ISL_496180\|2020-06-0 | ···················· | ···················· | ···················· | ···················· | ···C················ | ···················· |
| F | 4139 | hCoV-19/USA/WA-S1755/2020\|EPI_ISL_496181\|2020-06-0 | ···················· | ···················· | ···················· | ···················· | ···C················ | ···················· |
| F | 4140 | hCoV-19/USA/WA-S1756/2020\|EPI_ISL_496182\|2020-06-0 | ···················· | ···················· | ···················· | ···················· | ···················· | ···················· |
| F | 4141 | hCoV-19/USA/WA-S1757/2020\|EPI_ISL_496183\|2020-06-0 | ···················· | ···················· | ···················· | ···················· | ···················· | ···················· |
| F | 4142 | hCoV-19/USA/WA-S1758/2020\|EPI_ISL_496184\|2020-06-0 | ···················· | ···················· | ···················· | ···················· | ···················· | ···················· |
| F | 4143 | hCoV-19/USA/WA-S1760/2020\|EPI_ISL_496186\|2020-06-0 | ···················· | ···················· | ···················· | ···················· | ···················· | ···················· |
| F | 4144 | hCoV-19/USA/WA-S1761/2020\|EPI_ISL_496187\|2020-06-0 | ···················· | ···················· | ···················· | ···················· | ···················· | ···················· |
| F | 4145 | hCoV-19/USA/WA-S1762/2020\|EPI_ISL_496188\|2020-06-0 | ···················· | ···················· | ···················· | ···················· | ···················· | ···················· |
| F | 4146 | hCoV-19/USA/WA-S1764/2020\|EPI_ISL_496190\|2020-06-0 | ···················· | ···················· | ···················· | ···················· | ···················· | ···················· |
| F | 4147 | hCoV-19/USA/WA-S1765/2020\|EPI_ISL_496191\|2020-06-0 | ···················· | ···················· | ···················· | ···················· | ···················· | ···················· |
| F | 4148 | hCoV-19/USA/WA-S1766/2020\|EPI_ISL_496192\|2020-06-0 | ···················· | ···················· | ···················· | ···················· | ···················· | ···················· |
| F | 4149 | hCoV-19/USA/WA-S1767/2020\|EPI_ISL_496193\|2020-06-0 | ···················· | ···················· | ···················· | ···················· | ···················· | ···················· |
| F | 4150 | hCoV-19/USA/WA-S1768/2020\|EPI_ISL_496194\|2020-06-0 | ···················· | ···················· | ···················· | ···················· | ···················· | ···················· |
| F | 4151 | hCoV-19/USA/WA-S1769/2020\|EPI_ISL_496195\|2020-06-0 | ···················· | ···················· | ···················· | ···················· | ···················· | ···················· |
| F | 4152 | hCoV-19/USA/WA-S1770/2020\|EPI_ISL_496196\|2020-06-0 | ···················· | ···················· | ···················· | ···················· | ···················· | ···················· |
| F | 4153 | hCoV-19/USA/WA-S1771/2020\|EPI_ISL_496197\|2020-06-0 | ···················· | ···················· | ···················· | ···················· | ···················· | ···················· |
| F | 4154 | hCoV-19/USA/WA-S1773/2020\|EPI_ISL_496199\|2020-06-0 | ···················· | ···················· | ···················· | ···················· | ···················· | ···················· |
| F | 4155 | hCoV-19/USA/WA-S2016/2020\|EPI_ISL_497444\|2020-06-0 | ···················· | ···················· | ···················· | ···················· | ···················· | ···················· |
| F | 4156 | hCoV-19/USA/WA-S2017/2020\|EPI_ISL_497445\|2020-06-0 | ···················· | ···················· | ···················· | ···················· | ···················· | ···················· |
| F | 4157 | hCoV-19/USA/WA-S2018/2020\|EPI_ISL_497446\|2020-06-0 | ···················· | ···················· | ···················· | ···················· | ···················· | ···················· |
| F | 4158 | hCoV-19/USA/WA-S2019/2020\|EPI_ISL_497447\|2020-06-0 | ···················· | ···················· | ···················· | ···················· | ···················· | ···················· |
| F | 4159 | hCoV-19/USA/WA-S2020/2020\|EPI_ISL_497448\|2020-06-0 | ···················· | ···················· | ···················· | ···················· | ···················· | ···················· |
| F | 4160 | hCoV-19/USA/WA-S2021/2020\|EPI_ISL_497449\|2020-06-0 | ···················· | ···················· | ···················· | ···················· | ···················· | ···················· |
| F | 4161 | hCoV-19/USA/WA-S2022/2020\|EPI_ISL_497450\|2020-06-0 | ···················· | ···················· | ···················· | ···················· | ···················· | ···················· |
| F | 4162 | hCoV-19/USA/WA-S2025/2020\|EPI_ISL_497453\|2020-06-0 | ···················· | ···················· | ···················· | ···················· | ···················· | ···················· |
| F | 4163 | hCoV-19/USA/WA-S2026/2020\|EPI_ISL_497454\|2020-06-0 | ···················· | ···················· | ···················· | ···················· | ···················· | ···················· |
| F | 4164 | hCoV-19/USA/WA-S2027/2020\|EPI_ISL_497455\|2020-06-0 | ···················· | ···················· | ···················· | ···················· | ···················· | ···················· |
| F | 4165 | hCoV-19/USA/WA-S2028/2020\|EPI_ISL_497456\|2020-06-0 | ···················· | ···················· | ···················· | ···················· | ···················· | ···················· |
| F | 4166 | hCoV-19/USA/WA-S2029/2020\|EPI_ISL_497457\|2020-06-0 | ···················· | ···················· | ···················· | ···················· | ···················· | ···················· |
| F | 4167 | hCoV-19/USA/WA-S2030/2020\|EPI_ISL_497458\|2020-06-0 | ···················· | ···················· | ···················· | ···················· | ···················· | ···················· |
| F | 4168 | hCoV-19/USA/WA-S2031/2020\|EPI_ISL_497459\|2020-06-0 | ···················· | ···················· | ···················· | ···················· | ···················· | ···················· |
| F | 4169 | hCoV-19/USA/WA-S2032/2020\|EPI_ISL_497460\|2020-06-0 | ···················· | ···················· | ···················· | ···················· | ···················· | ···················· |
| F | 4170 | hCoV-19/USA/WA-S2033/2020\|EPI_ISL_497461\|2020-06-0 | ···················· | ···················· | ···················· | ···················· | ···················· | ···················· |
| F | 4171 | hCoV-19/USA/WA-S2034/2020\|EPI_ISL_497462\|2020-06-0 | ···················· | ···················· | ···················· | ···················· | ···················· | ···················· |
| F | 4172 | hCoV-19/USA/WA-S2035/2020\|EPI_ISL_497463\|2020-06-0 | ···················· | ···················· | ···················· | ···················· | ···················· | ···················· |
| F | 4173 | hCoV-19/USA/WA-S2036/2020\|EPI_ISL_497464\|2020-06-0 | ···················· | ···················· | ···················· | ···················· | ···················· | ···················· |
| F | 4174 | hCoV-19/USA/WA-S2037/2020\|EPI_ISL_497465\|2020-06-0 | ···················· | ···················· | ···················· | ···················· | ···················· | ···················· |
| F | 4175 | hCoV-19/USA/WA-S2060/2020\|EPI_ISL_497488\|2020-06-0 | ···················· | ···················· | ···················· | ···················· | ···················· | ···················· |
| F | 4176 | hCoV-19/USA/WA-S2061/2020\|EPI_ISL_497489\|2020-06-0 | ···················· | ···················· | ···················· | ···················· | ···················· | ···················· |
| F | 4177 | hCoV-19/USA/WA-S2062/2020\|EPI_ISL_497490\|2020-06-0 | ···················· | ···················· | ···················· | ···················· | ···················· | ···················· |
| F | 4178 | hCoV-19/USA/WA-S2063/2020\|EPI_ISL_497491\|2020-06-0 | ···················· | ···················· | ···················· | ···················· | ···················· | ···················· |
| F | 4179 | hCoV-19/USA/WA-S2064/2020\|EPI_ISL_497492\|2020-06-0 | ···················· | ···················· | ···················· | ···················· | ···················· | ···················· |
| F | 4180 | hCoV-19/USA/WA-S2065/2020\|EPI_ISL_497493\|2020-06-0 | ···················· | ···················· | ···················· | ···················· | ···················· | ···················· |
| F | 4181 | hCoV-19/USA/WA-S2066/2020\|EPI_ISL_497494\|2020-06-0 | ···················· | ···················· | ···················· | ···················· | ···················· | ···················· |
| F | 4182 | hCoV-19/USA/WA-S2067/2020\|EPI_ISL_497495\|2020-06-0 | ···················· | ···················· | ···················· | ···················· | ···················· | ···················· |
| F | 4183 | hCoV-19/USA/WA-S2068/2020\|EPI_ISL_497496\|2020-06-0 | ···················· | ···················· | ···················· | ···················· | ···················· | ···················· |

| | | | | | | | | |
|---|---|---|---|---|---|---|---|---|
| F | 4184 | hCoV-19/USA/WA-S2069/2020\|EPI_ISL_497497\|2020-06-0 | .................... | .................... | ..................... | .................... | .................... | .................... |
| F | 4185 | hCoV-19/USA/WA-S2070/2020\|EPI_ISL_497498\|2020-06-0 | .................... | .................... | ..................... | .................... | .................... | .................... |
| F | 4186 | hCoV-19/USA/WA-S2071/2020\|EPI_ISL_497499\|2020-06-0 | .................... | .................... | ..................... | .................... | .................... | .................... |
| F | 4187 | hCoV-19/USA/WA-S2277/2020\|EPI_ISL_497705\|2020-06-0 | .................... | .................... | ..................... | .................... | .................... | .................... |
| F | 4188 | hCoV-19/USA/WA-S2281/2020\|EPI_ISL_497709\|2020-06-1 | .................... | .................... | ..................... | .................... | .................... | .................... |
| F | 4189 | hCoV-19/Australia/NSW656/2020\|EPI_ISL_498775\|2020- | .................... | .................... | ..................... | .................... | .................... | .................... |
| F | 4190 | hCoV-19/Australia/NSW657/2020\|EPI_ISL_498776\|2020- | .................... | .................... | ..................... | .................... | .................... | .................... |
| F | 4191 | hCoV-19/Australia/NSW658/2020\|EPI_ISL_498777\|2020- | .................... | .................... | ..................... | .................... | .................... | .................... |
| F | 4192 | hCoV-19/Australia/NSW659/2020\|EPI_ISL_498778\|2020- | .................... | .................... | ..................... | .................... | .................... | .................... |
| F | 4193 | hCoV-19/Australia/NSW660/2020\|EPI_ISL_498779\|2020- | .................... | .................... | ..................... | .................... | .................... | .................... |
| F | 4194 | hCoV-19/Australia/NSW661/2020\|EPI_ISL_498780\|2020- | .................... | .................... | ..................... | .................... | .................... | .................... |
| F | 4195 | hCoV-19/Australia/NSW662/2020\|EPI_ISL_498781\|2020- | .................... | .................... | ..................... | .................... | .................... | .................... |
| F | 4196 | hCoV-19/Australia/NSW663/2020\|EPI_ISL_498782\|2020- | .................... | .................... | ..................... | .................... | .................... | .................... |
| F | 4197 | hCoV-19/Bangladesh/BCSIR-NILMRC-293/2020\|EPI_ISL_4 | .................... | .................... | ..................... | .................... | .................... | .................... |
| F | 4198 | hCoV-19/Bangladesh/BCSIR-NILMRC-297/2020\|EPI_ISL_4 | .................... | .................... | ..................... | .................... | .................... | .................... |
| F | 4199 | hCoV-19/Bangladesh/BCSIR-NILMRC-298/2020\|EPI_ISL_4 | .................... | .................... | ..................... | .................... | .................... | .................... |
| F | 4200 | hCoV-19/Bangladesh/BCSIR-NILMRC-306/2020\|EPI_ISL_4 | .................... | .................... | ..................... | .................... | .................... | .................... |
| F | 4201 | hCoV-19/Bangladesh/BCSIR-NILMRC-307/2020\|EPI_ISL_4 | .................... | .................... | ..................... | .................... | .................... | .................... |
| F | 4202 | hCoV-19/Bangladesh/BCSIR-NILMRC-315/2020\|EPI_ISL_4 | .................... | .................... | ..................... | .................... | .................... | .................... |
| F | 4203 | hCoV-19/Bangladesh/BCSIR-NILMRC-318/2020\|EPI_ISL_4 | .................... | .................... | ..................... | .................... | .................... | .................... |
| F | 4204 | hCoV-19/Bangladesh/BCSIR-NILMRC-322/2020\|EPI_ISL_4 | .................... | .................... | ..................... | .................... | .................... | .................... |
| F | 4205 | hCoV-19/Bangladesh/BCSIR-NILMRC-323/2020\|EPI_ISL_4 | .................... | .................... | ..................... | .................... | .................... | .................... |
| F | 4206 | hCoV-19/Bangladesh/BCSIR-NILMRC-326/2020\|EPI_ISL_4 | .................... | .................... | ..................... | .................... | .................... | .................... |
| F | 4207 | hCoV-19/Bangladesh/BCSIR-NILMRC-327/2020\|EPI_ISL_4 | .................... | .................... | ..................... | .................... | .................... | .................... |
| F | 4208 | hCoV-19/Bangladesh/BCSIR-NILMRC-328/2020\|EPI_ISL_4 | .................... | .................... | ..................... | .................... | .................... | .................... |
| F | 4209 | hCoV-19/Bangladesh/BCSIR-NILMRC-329/2020\|EPI_ISL_4 | .................... | .................... | ..................... | .................... | .................... | .................... |
| F | 4210 | hCoV-19/Bangladesh/BCSIR-NILMRC-333/2020\|EPI_ISL_4 | .................... | .................... | ..................... | .................... | .................... | .................... |
| F | 4213 | hCoV-19/England/LIVE-A4A97/2020\|EPI_ISL_500067\|202 | .................... | .................... | ..................... | .................... | .................... | .................... |
| F | 4214 | hCoV-19/England/LIVE-A4AB5/2020\|EPI_ISL_500068\|202 | .................... | .................... | ..................... | .................... | .................... | .................... |
| F | 4215 | hCoV-19/Greece/14999_HPI/2020\|EPI_ISL_501248\|2020- | .................... | .................... | ..................... | .................... | .................... | .................... |
| F | 4216 | hCoV-19/Greece/15005_HPI/2020\|EPI_ISL_501249\|2020- | .................... | .................... | ..................... | .................... | .................... | .................... |
| F | 4217 | hCoV-19/Greece/22565_HPI/2020\|EPI_ISL_501250\|2020- | .................... | .................... | ..................... | .................... | .................... | .................... |
| F | 4218 | hCoV-19/Greece/8334_HPI/2020\|EPI_ISL_501251\|2020-0 | .................... | .................... | ..................... | .................... | .................... | .................... |
| F | 4219 | hCoV-19/Ireland/LH-NVRL-20IRL22956/2020\|EPI_ISL_50 | .................... | .................... | ..................... | .................... | .................... | .................... |
| F | 4221 | hCoV-19/Germany/BW-UT-011/2020\|EPI_ISL_508690\|2020 | .................... | .................... | ..................... | .................... | .................... | .................... |
| F | 4222 | hCoV-19/Germany/BW-UT-012/2020\|EPI_ISL_508691\|2020 | .................... | .................... | ..................... | .................... | .................... | .................... |
| F | 4223 | hCoV-19/Germany/BW-UT-014/2020\|EPI_ISL_508693\|2020 | .................... | .................... | ..................... | .................... | .................... | .................... |
| F | 4224 | hCoV-19/Germany/BW-UT-015/2020\|EPI_ISL_508694\|2020 | .................... | .................... | ..................... | .................... | .................... | .................... |
| F | 4225 | hCoV-19/Germany/BW-UT-018/2020\|EPI_ISL_508696\|2020 | .................... | .................... | ..................... | .................... | .................... | .................... |
| F | 4226 | hCoV-19/Germany/BW-UT-019/2020\|EPI_ISL_508697\|2020 | .................... | .................... | ..................... | .................... | .................... | .................... |
| F | 4227 | hCoV-19/Germany/BW-UT-024/2020\|EPI_ISL_508699\|2020 | .................... | .................... | ..................... | .................... | .................... | .................... |
| F | 4228 | hCoV-19/Madagascar/IP-01650/2020\|EPI_ISL_508862\|20 | .................... | .................... | ..................... | .................... | .................... | .................... |
| F | 4229 | hCoV-19/Madagascar/IP-01861/2020\|EPI_ISL_508863\|20 | .................... | .................... | ..................... | .................... | .................... | .................... |
| F | 4230 | hCoV-19/Switzerland/ZH-UZH-1000482982/2020\|EPI_ISL | .................... | .................... | ..................... | .................... | .................... | .................... |
| F | 4231 | hCoV-19/Switzerland/ZH-UZH-1000482983/2020\|EPI_ISL | .................... | .................... | ..................... | .................... | .................... | .................... |
| F | 4232 | hCoV-19/Switzerland/ZH-UZH-1000482984/2020\|EPI_ISL | .................... | .................... | ..................... | .................... | .................... | .................... |
| F | 4233 | hCoV-19/Switzerland/ZH-UZH-1000482985/2020\|EPI_ISL | .................... | .................... | ..................... | .................... | .................... | .................... |
| F | 4234 | hCoV-19/Switzerland/ZH-UZH-1000483258/2020\|EPI_ISL | .................... | .................... | ..................... | .................... | .................... | .................... |
| F | 4235 | hCoV-19/Switzerland/ZH-UZH-1000484569/2020\|EPI_ISL | .................... | .................... | ..................... | .................... | .................... | .................... |
| F | 4236 | hCoV-19/Switzerland/ZH-UZH-1000486784/2020\|EPI_ISL | .................... | .................... | ..................... | .................... | .................... | .................... |
| F | 4237 | hCoV-19/France/ARA-01890/2020\|EPI_ISL_508871\|2020- | .................... | .................... | ..................... | .................... | .................... | .................... |
| F | 4238 | hCoV-19/France/ARA-07639/2020\|EPI_ISL_508872\|2020- | .................... | .................... | ..................... | .................... | .................... | .................... |
| F | 4239 | hCoV-19/France/ARA-07910/2020\|EPI_ISL_508873\|2020- | .................... | .................... | ..................... | .................... | .................... | .................... |
| F | 4240 | hCoV-19/France/ARA-08589/2020\|EPI_ISL_508874\|2020- | .................... | .................... | ..................... | .................... | .................... | .................... |
| F | 4241 | hCoV-19/France/ARA-0873/2020\|EPI_ISL_508875\|2020-0 | .................... | .................... | ..................... | .................... | .................... | .................... |
| F | 4242 | hCoV-19/France/ARA-08938/2020\|EPI_ISL_508876\|2020- | .................... | .................... | ..................... | .................... | .................... | .................... |
| F | 4243 | hCoV-19/France/ARA-09178/2020\|EPI_ISL_508877\|2020- | .................... | .................... | ..................... | .................... | .................... | .................... |
| F | 4244 | hCoV-19/France/ARA-09193/2020\|EPI_ISL_508878\|2020- | .................... | .................... | ..................... | .................... | .................... | .................... |
| F | 4245 | hCoV-19/France/ARA-10160/2020\|EPI_ISL_508879\|2020- | .................... | .................... | ..................... | .................... | .................... | .................... |
| F | 4246 | hCoV-19/France/ARA-10175/2020\|EPI_ISL_508880\|2020- | .................... | .................... | ..................... | .................... | .................... | .................... |
| F | 4247 | hCoV-19/France/ARA-10242/2020\|EPI_ISL_508881\|2020- | .................... | .................... | ..................... | .................... | .................... | .................... |
| F | 4248 | hCoV-19/France/ARA-10823/2020\|EPI_ISL_508882\|2020- | .................... | .................... | ..................... | .................... | .................... | .................... |
| F | 4249 | hCoV-19/France/ARA-10825/2020\|EPI_ISL_508883\|2020- | .................... | .................... | ..................... | .................... | .................... | .................... |
| F | 4250 | hCoV-19/France/ARA-14226/2020\|EPI_ISL_508884\|2020- | .................... | .................... | ..................... | .................... | .................... | .................... |
| F | 4251 | hCoV-19/France/ARA-14350/2020\|EPI_ISL_508885\|2020- | .................... | .................... | ..................... | .................... | .................... | .................... |
| F | 4252 | hCoV-19/France/ARA-14353/2020\|EPI_ISL_508886\|2020- | .................... | .................... | ..................... | .................... | .................... | .................... |
| F | 4253 | hCoV-19/France/ARA-14443/2020\|EPI_ISL_508887\|2020- | .................... | .................... | ..................... | .................... | .................... | .................... |
| F | 4254 | hCoV-19/France/ARA-14456/2020\|EPI_ISL_508888\|2020- | .................... | .................... | ..................... | .................... | .................... | .................... |
| F | 4255 | hCoV-19/France/ARA-14570/2020\|EPI_ISL_508889\|2020- | .................... | .................... | ..................... | .................... | .................... | .................... |
| F | 4256 | hCoV-19/France/ARA-14623/2020\|EPI_ISL_508890\|2020- | .................... | .................... | ..................... | .................... | .................... | .................... |
| F | 4257 | hCoV-19/France/ARA-14624/2020\|EPI_ISL_508891\|2020- | .................... | .................... | ..................... | .................... | .................... | .................... |
| F | 4258 | hCoV-19/France/ARA-14627/2020\|EPI_ISL_508892\|2020- | .................... | .................... | ..................... | .................... | .................... | .................... |
| F | 4259 | hCoV-19/France/ARA-14735/2020\|EPI_ISL_508893\|2020- | .................... | .................... | ..................... | .................... | .................... | .................... |
| F | 4260 | hCoV-19/France/ARA-14791/2020\|EPI_ISL_508894\|2020- | .................... | .................... | ..................... | .................... | .................... | .................... |
| F | 4261 | hCoV-19/France/ARA-1492/2020\|EPI_ISL_508895\|2020-0 | .................... | .................... | ..................... | .................... | .................... | .................... |
| F | 4262 | hCoV-19/France/ARA-14951/2020\|EPI_ISL_508896\|2020- | .................... | .................... | ..................... | .................... | .................... | .................... |
| F | 4263 | hCoV-19/France/ARA-14953/2020\|EPI_ISL_508897\|2020- | .................... | .................... | ..................... | .................... | .................... | .................... |
| F | 4264 | hCoV-19/France/ARA-14993/2020\|EPI_ISL_508898\|2020- | .................... | .................... | ..................... | .................... | .................... | .................... |
| F | 4265 | hCoV-19/France/ARA-14997/2020\|EPI_ISL_508899\|2020- | .................... | .................... | ..................... | .................... | .................... | .................... |
| F | 4266 | hCoV-19/Switzerland/VD-ETHZ-220060/2020\|EPI_ISL_51 | .................... | .................... | ..................... | .................... | .................... | .................... |

| | | | | | | | |
|---|---|---|---|---|---|---|---|
| F | 4267 hCoV-19/Switzerland/BE-ETHZ-220062/2020\|EPI_ISL_51 | ···················· | ·A·················· | ···················· | ···················· | ···················· | ···················· |
| F | 4268 hCoV-19/Switzerland/AG-ETHZ-220063/2020\|EPI_ISL_51 | ···················· | ···················· | ···················· | ···················· | ···················· | ···················· |
| F | 4269 hCoV-19/Switzerland/BL-ETHZ-220064/2020\|EPI_ISL_51 | ···················· | ···················· | ···················· | ···················· | ···················· | ···················· |
| F | 4270 hCoV-19/Switzerland/BE-ETHZ-220065/2020\|EPI_ISL_51 | ···················· | ···················· | ···················· | ···················· | ···················· | ···················· |
| F | 4271 hCoV-19/Switzerland/VD-ETHZ-220066/2020\|EPI_ISL_51 | ···················· | ···················· | ···················· | ···················· | ···················· | ···················· |
| F | 4272 hCoV-19/Switzerland/BL-ETHZ-220067/2020\|EPI_ISL_51 | ···················· | ···················· | ···················· | ···················· | ···················· | ···················· |
| F | 4273 hCoV-19/Switzerland/BE-ETHZ-220068/2020\|EPI_ISL_51 | ···················· | ······A············· | ···················· | ···················· | ···················· | ···················· |
| F | 4274 hCoV-19/Switzerland/ZH-ETHZ-220069/2020\|EPI_ISL_51 | ···················· | ···················· | ···················· | ···················· | ···················· | ···················· |
| F | 4275 hCoV-19/Switzerland/ZH-ETHZ-220070/2020\|EPI_ISL_51 | ···················· | ···················· | ···················· | ···················· | ···················· | ···················· |
| F | 4276 hCoV-19/SaudiArabia/KAUST-MAKKAH295/2020\|EPI_ISL_5 | ···················· | ···················· | ···················· | ···················· | ···················· | ···················· |
| F | 4277 hCoV-19/SaudiArabia/KAUST-MAKKAH297/2020\|EPI_ISL_5 | ···················· | ·A·················· | ···················· | ···················· | ···················· | ···A················ |
| F | 4278 hCoV-19/SaudiArabia/KAUST-MAKKAH300/2020\|EPI_ISL_5 | ···················· | ···················· | ···················· | ···················· | ···················· | ···················· |
| F | 4279 hCoV-19/SaudiArabia/KAUST-MAKKAH301/2020\|EPI_ISL_5 | ···················· | ·A·················· | ···················· | ···················· | ···················· | ···A················ |
| F | 4280 hCoV-19/SaudiArabia/KAUST-MAKKAH303/2020\|EPI_ISL_5 | ···················· | ···················· | ···················· | ···················· | ···················· | ···A················ |
| F | 4281 hCoV-19/SaudiArabia/KAUST-MAKKAH309/2020\|EPI_ISL_5 | ···················· | ···················· | ···················· | ···················· | ···················· | ···················· |
| F | 4282 hCoV-19/Saudi Arabia/KAUST-MAKKAH726/2020\|EPI_ISL_ | ···················· | ···················· | ···················· | ···················· | ···················· | ···················· |
| F | 4283 hCoV-19/Saudi Arabia/KAUST-MAKKAH727/2020\|EPI_ISL_ | ···················· | ···················· | ···················· | ···················· | ···················· | ···················· |
| F | 4284 hCoV-19/Saudi Arabia/KAUST-MAKKAH730/2020\|EPI_ISL_ | ···················· | ···················· | ···················· | ···················· | ···················· | ···················· |
| F | 4285 hCoV-19/Saudi Arabia/KAUST-MAKKAH735/2020\|EPI_ISL_ | ···················· | ···················· | ···················· | ···················· | ···················· | ···················· |
| F | 4286 hCoV-19/Saudi Arabia/KAUST-MAKKAH741/2020\|EPI_ISL_ | ···················· | ···················· | ···················· | ···················· | ···················· | ···················· |
| F | 4287 hCoV-19/SaudiArabia/KAUST-MAKKAH760/2020\|EPI_ISL_5 | ···················· | ···················· | ···················· | ···················· | ···················· | ···················· |
| F | 4288 hCoV-19/SaudiArabia/KAUST-MAKKAH761/2020\|EPI_ISL_5 | ···················· | ·A·················· | ···················· | ···················· | ···················· | ···A················ |
| F | 4289 hCoV-19/SaudiArabia/KAUST-MAKKAH762/2020\|EPI_ISL_5 | ···················· | ···················· | ···················· | ···················· | ···················· | ···················· |
| F | 4290 hCoV-19/SaudiArabia/KAUST-MAKKAH768/2020\|EPI_ISL_5 | ···················· | ···················· | ···················· | ···················· | ···················· | ···················· |
| F | 4291 hCoV-19/Australia/NSW717/2020\|EPI_ISL_513360\|2020- | ···················· | ···················· | ···················· | ···················· | ···················· | ···················· |
| F | 4292 hCoV-19/Australia/NSW718/2020\|EPI_ISL_513361\|2020- | ···················· | ···················· | ···················· | ···················· | ···················· | ···················· |
| F | 4293 hCoV-19/Australia/NSW719/2020\|EPI_ISL_513362\|2020- | ···················· | ···················· | ···················· | ···················· | ···················· | ···················· |
| F | 4294 hCoV-19/Australia/NSW720/2020\|EPI_ISL_513363\|2020- | ···················· | ···················· | ···················· | ···················· | ···················· | ···················· |
| F | 4295 hCoV-19/Australia/NSW721/2020\|EPI_ISL_513364\|2020- | ···················· | ···················· | ···················· | ···················· | ···················· | ···················· |
| F | 4296 hCoV-19/Australia/NSW722/2020\|EPI_ISL_513365\|2020- | ···················· | ···················· | ···················· | ···················· | ···················· | ···················· |
| F | 4297 hCoV-19/Australia/NSW723/2020\|EPI_ISL_513366\|2020- | ···················· | ···················· | ···················· | ···················· | ···················· | ···················· |
| F | 4298 hCoV-19/Australia/NSW724/2020\|EPI_ISL_513367\|2020- | ···················· | ···················· | ···················· | ···················· | ···················· | ···················· |
| F | 4299 hCoV-19/Australia/NSW725/2020\|EPI_ISL_513368\|2020- | ···················· | ···················· | ···················· | ···················· | ···················· | ···················· |
| F | 4300 hCoV-19/Australia/NSW726/2020\|EPI_ISL_513369\|2020- | ···················· | ···················· | ···················· | ···················· | ···················· | ···················· |
| F | 4301 hCoV-19/Scotland/EDB7140/2020\|EPI_ISL_514549\|2020- | ···················· | ···················· | ···················· | ···················· | ···················· | ···················· |
| F | 4302 hCoV-19/Scotland/EDB7142/2020\|EPI_ISL_514550\|2020- | ···················· | ···················· | ···················· | ···················· | ···················· | ···················· |
| F | 4303 hCoV-19/Scotland/EDB7145/2020\|EPI_ISL_514551\|2020- | ···················· | ···················· | ···················· | ···················· | ···················· | ···················· |
| F | 4304 hCoV-19/Scotland/EDB7146/2020\|EPI_ISL_514552\|2020- | ···················· | ···················· | ···················· | ···················· | ···················· | ···················· |
| F | 4305 hCoV-19/Scotland/EDB7147/2020\|EPI_ISL_514553\|2020- | ···················· | ···················· | ···················· | ···················· | ···················· | ···················· |
| F | 4306 hCoV-19/Scotland/EDB7148/2020\|EPI_ISL_514554\|2020- | ···················· | ···················· | ···················· | ···················· | ···················· | ···················· |
| F | 4307 hCoV-19/Scotland/EDB7149/2020\|EPI_ISL_514555\|2020- | ···················· | ···················· | ···················· | ···················· | ···················· | ···················· |
| F | 4308 hCoV-19/Scotland/EDB7151/2020\|EPI_ISL_514557\|2020- | ···················· | ···················· | ···················· | ···················· | ···················· | ···················· |
| F | 4309 hCoV-19/Scotland/EDB7152/2020\|EPI_ISL_514558\|2020- | ···················· | ···················· | ···················· | ···················· | ···················· | ···················· |
| F | 4310 hCoV-19/Scotland/EDB7153/2020\|EPI_ISL_514559\|2020- | ···················· | ···················· | ···················· | ···················· | ···················· | ···················· |
| F | 4311 hCoV-19/Scotland/EDB7154/2020\|EPI_ISL_514560\|2020- | ···················· | ···················· | ···················· | ···················· | ···················· | ···················· |
| F | 4312 hCoV-19/Scotland/EDB7155/2020\|EPI_ISL_514561\|2020- | ···················· | ···················· | ···················· | ···················· | ···················· | ···················· |
| F | 4313 hCoV-19/Scotland/EDB7156/2020\|EPI_ISL_514562\|2020- | ···················· | ···················· | ···················· | ···················· | ···················· | ···················· |
| F | 4314 hCoV-19/Wales/PHWC-1699EB/2020\|EPI_ISL_514563\|2020 | ···················· | ···················· | ···················· | ···················· | ···················· | ···················· |
| F | 4315 hCoV-19/Wales/PHWC-1699FA/2020\|EPI_ISL_514564\|2020 | ···················· | ···················· | ···················· | ···················· | ···················· | ···················· |
| F | 4316 hCoV-19/Wales/PHWC-169A06/2020\|EPI_ISL_514565\|2020 | ···················· | ···················· | ···················· | ···················· | ···················· | ···················· |
| F | 4317 hCoV-19/Wales/PHWC-169A15/2020\|EPI_ISL_514566\|2020 | ···················· | ···················· | ···················· | ···················· | ···················· | ···················· |
| F | 4318 hCoV-19/Wales/PHWC-169A24/2020\|EPI_ISL_514567\|2020 | ···················· | ···················· | ···················· | ···················· | ···················· | ···················· |
| F | 4319 hCoV-19/Wales/PHWC-169A33/2020\|EPI_ISL_514568\|2020 | ···················· | ···················· | ···················· | ···················· | ···················· | ···················· |
| F | 4320 hCoV-19/Wales/PHWC-169A42/2020\|EPI_ISL_514569\|2020 | ···················· | ···················· | ···················· | ···················· | ···················· | ···················· |
| F | 4321 hCoV-19/South Africa/KRISP-K001122/2020\|EPI_ISL_51 | ···················· | ···················· | ···················· | ···················· | ···················· | ···················· |
| F | 4322 hCoV-19/South Africa/KRISP-K001317/2020\|EPI_ISL_51 | ···················· | ···················· | ···················· | ···················· | ···················· | ···················· |
| F | 4323 hCoV-19/South Africa/KRISP-K001318/2020\|EPI_ISL_51 | ···················· | ···················· | ···················· | ···················· | ···················· | ···················· |
| F | 4324 hCoV-19/South Africa/KRISP-K001319/2020\|EPI_ISL_51 | ···················· | ···················· | ···················· | ···················· | ···················· | ···················· |
| F | 4325 hCoV-19/South Africa/KRISP-K001320/2020\|EPI_ISL_51 | ···················· | ···················· | ···················· | ···················· | ···················· | ···················· |
| F | 4326 hCoV-19/South Africa/KRISP-K001321/2020\|EPI_ISL_51 | ···················· | ···················· | ···················· | ···················· | ···················· | ···················· |
| F | 4327 hCoV-19/South Africa/KRISP-K001322/2020\|EPI_ISL_51 | ···················· | ···················· | ···················· | ···················· | ···················· | ···················· |
| F | 4328 hCoV-19/South Africa/KRISP-K001323/2020\|EPI_ISL_51 | ···················· | ···················· | ···················· | ···················· | ···················· | ···················· |
| F | 4329 hCoV-19/South Africa/KRISP-K001324/2020\|EPI_ISL_51 | ···················· | ···················· | ···················· | ···················· | ···················· | ···················· |
| F | 4330 hCoV-19/South Africa/KRISP-K001325/2020\|EPI_ISL_51 | ···················· | ···················· | ···················· | ···················· | ···················· | ···················· |
| F | 4331 hCoV-19/South Africa/KRISP-K001326/2020\|EPI_ISL_51 | ···················· | ···················· | ···················· | ···················· | ···················· | ···················· |
| F | 4332 hCoV-19/South Africa/KRISP-K001327/2020\|EPI_ISL_51 | ···················· | ···················· | ···················· | ···················· | ···················· | ···················· |
| F | 4333 hCoV-19/South Africa/KRISP-K001328/2020\|EPI_ISL_51 | ···················· | ···················· | ···················· | ···················· | ···················· | ···················· |
| F | 4334 hCoV-19/South Africa/KRISP-K001397/2020\|EPI_ISL_51 | ···················· | ···················· | ···················· | ···················· | ···················· | ···················· |
| F | 4335 hCoV-19/South Africa/KRISP-K001398/2020\|EPI_ISL_51 | ········T··········· | ···················· | ···················· | ···················· | ···················· | ···················· |
| F | 4336 hCoV-19/South Africa/KRISP-K001399/2020\|EPI_ISL_51 | ···················· | ···················· | ···················· | ···················· | ···················· | ···················· |
| F | 4337 hCoV-19/South Africa/KRISP-K001400/2020\|EPI_ISL_51 | ···················· | ···················· | ···················· | ···················· | ···················· | ···················· |
| F | 4338 hCoV-19/South Africa/KRISP-K001401/2020\|EPI_ISL_51 | ···················· | ···················· | ···················· | ···················· | ···················· | ···················· |
| F | 4339 hCoV-19/South Africa/KRISP-K001402/2020\|EPI_ISL_51 | ···················· | ···················· | ···················· | ···················· | ···················· | ···················· |
| F | 4340 hCoV-19/South Africa/KRISP-K001403/2020\|EPI_ISL_51 | ···················· | ···················· | ···················· | ···················· | ···················· | ···················· |
| F | 4341 hCoV-19/South Africa/KRISP-K001404/2020\|EPI_ISL_51 | ···················· | ···················· | ···················· | ···················· | ···················· | ···················· |
| F | 4342 hCoV-19/South Africa/KRISP-K001405/2020\|EPI_ISL_51 | ···················· | ···················· | ···················· | ···················· | ···················· | ···················· |
| F | 4343 hCoV-19/South Africa/KRISP-K001406/2020\|EPI_ISL_51 | ···················· | ···················· | ···················· | ···················· | ···················· | ···················· |
| F | 4344 hCoV-19/South Africa/KRISP-K001407/2020\|EPI_ISL_51 | ···················· | ···················· | ···················· | ···················· | ···················· | ···················· |
| F | 4345 hCoV-19/South Africa/KRISP-K001408/2020\|EPI_ISL_51 | ···················· | ···················· | ···················· | ···················· | ···················· | ···················· |
| F | 4346 hCoV-19/South Africa/KRISP-K001409/2020\|EPI_ISL_51 | ···················· | ···················· | ···················· | ···················· | ···················· | ···················· |

| | | | | | | | | |
|---|---|---|---|---|---|---|---|---|
| F | 4347 | hCoV-19/South Africa/KRISP-K001410/2020\|EPI_ISL_51 | .................... | .................... | ..................... | .................... | .................... | .................... |
| F | 4348 | hCoV-19/USA/CA-CDPH001/2020\|EPI_ISL_515894\|2020-03 | .................... | .................... | ..................... | .................... | .................... | .................... |
| F | 4349 | hCoV-19/USA/CA-CDPH003/2020\|EPI_ISL_515896\|2020-03 | .................... | .................... | ..................... | .................... | .................... | .................... |
| F | 4350 | hCoV-19/USA/CA-CDPH004/2020\|EPI_ISL_515897\|2020-03 | .................... | .................... | ..................... | .................... | .................... | .................... |
| F | 4351 | hCoV-19/USA/CA-CDPH005/2020\|EPI_ISL_515898\|2020-03 | .................... | .................... | ..................... | .................... | .................... | .................... |
| F | 4352 | hCoV-19/USA/CA-CDPH006/2020\|EPI_ISL_515899\|2020-03 | .................... | .................... | ..................... | .................... | .................... | .................... |
| F | 4353 | hCoV-19/USA/WA-S1676/2020\|EPI_ISL_496102\|2020-05-2 | .................... | .................... | ..................... | .................... | .................... | .................... |
| F | 4354 | hCoV-19/USA/WA-S1677/2020\|EPI_ISL_496103\|2020-05-2 | .................... | .................... | ..................... | .................... | .................... | .................... |
| F | 4355 | hCoV-19/USA/WA-S1678/2020\|EPI_ISL_496104\|2020-05-2 | .................... | .................... | ..................... | .................... | .................... | .................... |
| F | 4356 | hCoV-19/USA/WA-S1679/2020\|EPI_ISL_496105\|2020-05-2 | .................... | .................... | ..................... | .................... | .................... | .................... |
| F | 4357 | hCoV-19/USA/WA-S1680/2020\|EPI_ISL_496106\|2020-05-2 | .................... | .................... | ..................... | .................... | .................... | .................... |
| F | 4358 | hCoV-19/USA/WA-S1681/2020\|EPI_ISL_496107\|2020-05-2 | .................... | .................... | ..................... | .................... | .................... | .................... |
| F | 4359 | hCoV-19/USA/WA-S1682/2020\|EPI_ISL_496108\|2020-05-2 | .................... | .................... | ..................... | .................... | .................... | .................... |
| F | 4360 | hCoV-19/USA/WA-S1685/2020\|EPI_ISL_496111\|2020-05-2 | .................... | .................... | ..................... | .................... | .................... | .................... |
| F | 4361 | hCoV-19/USA/WA-S1686/2020\|EPI_ISL_496112\|2020-05-3 | .................... | .................... | ..................... | .................... | .................... | .................... |
| F | 4362 | hCoV-19/USA/WA-S1695/2020\|EPI_ISL_496121\|2020-05-3 | .................... | .................... | ..................... | .................... | .................... | .................... |
| F | 4363 | hCoV-19/USA/WA-S1709/2020\|EPI_ISL_496135\|2020-06-0 | .................... | .................... | ..................... | .................... | .................... | .................... |
| F | 4364 | hCoV-19/USA/WA-S1710/2020\|EPI_ISL_496136\|2020-05-3 | .................... | .................... | ..................... | .................... | .................... | .................... |
| F | 4365 | hCoV-19/USA/WA-S1711/2020\|EPI_ISL_496137\|2020-06-0 | .................... | .................... | ..................... | .................... | .................... | .................... |
| F | 4366 | hCoV-19/USA/WA-S1712/2020\|EPI_ISL_496138\|2020-06-0 | .................... | .................... | ..................... | .................... | .................... | .................... |
| F | 4367 | hCoV-19/USA/WA-S1713/2020\|EPI_ISL_496139\|2020-06-0 | .................... | .................... | ..................... | .................... | .................... | .................... |
| F | 4368 | hCoV-19/USA/WA-S1714/2020\|EPI_ISL_496140\|2020-06-0 | .................... | .................... | ..................... | .................... | .................... | .................... |
| F | 4369 | hCoV-19/USA/WA-S1715/2020\|EPI_ISL_496141\|2020-06-0 | .................... | .................... | ..................... | .................... | .................... | .................... |
| F | 4370 | hCoV-19/USA/WA-S1716/2020\|EPI_ISL_496142\|2020-06-0 | .................... | .................... | ..................... | .................... | .................... | .................... |
| F | 4371 | hCoV-19/USA/WA-S1717/2020\|EPI_ISL_496143\|2020-06-0 | .................... | .................... | ..................... | .................... | .................... | .................... |
| F | 4372 | hCoV-19/USA/WA-S1718/2020\|EPI_ISL_496144\|2020-06-0 | .................... | .................... | ..................... | .................... | .................... | .................... |
| F | 4373 | hCoV-19/USA/WA-S1719/2020\|EPI_ISL_496145\|2020-06-0 | .................... | .................... | ..................... | .................... | .................... | .................... |
| F | 4374 | hCoV-19/USA/WA-S1720/2020\|EPI_ISL_496146\|2020-06-0 | .................... | .................... | ..................... | .................... | .................... | .................... |
| F | 4375 | hCoV-19/USA/WA-S1721/2020\|EPI_ISL_496147\|2020-06-0 | .................... | .................... | ..................... | .................... | .................... | .................... |
| F | 4376 | hCoV-19/USA/WA-S1722/2020\|EPI_ISL_496148\|2020-06-0 | .................... | .................... | ..................... | .................... | .................... | .................... |
| F | 4377 | hCoV-19/USA/WA-S1723/2020\|EPI_ISL_496149\|2020-06-0 | .................... | .................... | ..................... | .................... | .................... | .................... |
| F | 4378 | hCoV-19/USA/WA-S1724/2020\|EPI_ISL_496150\|2020-06-0 | .................... | .................... | ..................... | .................... | .................... | .................... |
| F | 4379 | hCoV-19/USA/WA-S1725/2020\|EPI_ISL_496151\|2020-06-0 | .................... | .................... | ..................... | .................... | .................... | .................... |
| F | 4380 | hCoV-19/USA/WA-S1726/2020\|EPI_ISL_496152\|2020-06-0 | .................... | .................... | ..................... | .................... | .................... | .................... |
| F | 4381 | hCoV-19/USA/WA-S1727/2020\|EPI_ISL_496153\|2020-06-0 | .................... | .................... | ..................... | .................... | .................... | .................... |
| F | 4382 | hCoV-19/USA/WA-S1728/2020\|EPI_ISL_496154\|2020-06-0 | .................... | .................... | ..................... | .................... | .................... | .................... |
| F | 4383 | hCoV-19/USA/WA-S1729/2020\|EPI_ISL_496155\|2020-06-0 | .................... | .................... | ..................... | .................... | .................... | .................... |
| F | 4384 | hCoV-19/USA/WA-S1730/2020\|EPI_ISL_496156\|2020-06-0 | .................... | .................... | ..................... | .................... | .................... | .................... |
| F | 4385 | hCoV-19/USA/WA-S1972/2020\|EPI_ISL_497400\|2020-06-0 | .................... | .................... | ..................... | .................... | .................... | .................... |
| F | 4386 | hCoV-19/USA/WA-S1973/2020\|EPI_ISL_497401\|2020-06-0 | .................... | .................... | ..................... | .................... | .................... | .................... |
| F | 4387 | hCoV-19/USA/WA-S1974/2020\|EPI_ISL_497402\|2020-06-0 | .................... | .................... | ..................... | .................... | .................... | .................... |
| F | 4388 | hCoV-19/USA/WA-S1975/2020\|EPI_ISL_497403\|2020-06-0 | .................... | .................... | ..................... | .................... | .................... | .................... |
| F | 4389 | hCoV-19/USA/WA-S1976/2020\|EPI_ISL_497404\|2020-06-0 | .................... | .................... | ..................... | .................... | .................... | .................... |
| F | 4390 | hCoV-19/USA/WA-S1977/2020\|EPI_ISL_497405\|2020-06-0 | .................... | .................... | ..................... | .................... | .................... | .................... |
| F | 4391 | hCoV-19/USA/WA-S1978/2020\|EPI_ISL_497406\|2020-06-0 | .................... | .................... | ..................... | .................... | .................... | .................... |
| F | 4392 | hCoV-19/USA/WA-S1979/2020\|EPI_ISL_497407\|2020-06-0 | .................... | .................... | ..................... | .................... | .................... | .................... |
| F | 4393 | hCoV-19/USA/WA-S1980/2020\|EPI_ISL_497408\|2020-06-0 | .................... | .................... | ..................... | .................... | .................... | .................... |
| F | 4394 | hCoV-19/USA/WA-S1981/2020\|EPI_ISL_497409\|2020-06-0 | .................... | .................... | ..................... | .................... | .................... | .................... |
| F | 4395 | hCoV-19/USA/WA-S1982/2020\|EPI_ISL_497410\|2020-06-0 | .................... | .................... | ..................... | .................... | .................... | .................... |
| F | 4396 | hCoV-19/USA/WA-S1983/2020\|EPI_ISL_497411\|2020-06-0 | .................... | .................... | ..................... | .................... | .................... | .................... |
| F | 4397 | hCoV-19/USA/WA-S1984/2020\|EPI_ISL_497412\|2020-06-0 | .................... | .................... | ..................... | .................... | .................... | .................... |
| F | 4398 | hCoV-19/USA/WA-S1985/2020\|EPI_ISL_497413\|2020-06-0 | .................... | .................... | ..................... | .................... | .................... | .................... |
| F | 4399 | hCoV-19/USA/WA-S1986/2020\|EPI_ISL_497414\|2020-06-0 | .................... | .................... | ..................... | .................... | .................... | .................... |
| F | 4400 | hCoV-19/USA/WA-S1987/2020\|EPI_ISL_497415\|2020-06-0 | .................... | .................... | ..................... | .................... | .................... | .................... |
| F | 4401 | hCoV-19/USA/WA-S1988/2020\|EPI_ISL_497416\|2020-06-0 | .................... | .................... | ..................... | .................... | .................... | .................... |
| F | 4402 | hCoV-19/USA/WA-S1989/2020\|EPI_ISL_497417\|2020-06-0 | .................... | .................... | ..................... | .................... | .................... | .................... |
| F | 4403 | hCoV-19/USA/WA-S1990/2020\|EPI_ISL_497418\|2020-06-0 | .................... | .................... | ..................... | .................... | .................... | .................... |
| F | 4404 | hCoV-19/USA/WA-S1992/2020\|EPI_ISL_497420\|2020-06-0 | .................... | .................... | ..................... | .................... | .................... | .................... |
| F | 4405 | hCoV-19/USA/WA-S1993/2020\|EPI_ISL_497421\|2020-06-0 | .................... | .................... | ..................... | .................... | .................... | .................... |
| F | 4406 | hCoV-19/USA/WA-S2005/2020\|EPI_ISL_497433\|2020-06-0 | .................... | .................... | ..................... | .................... | .................... | .................... |
| F | 4407 | hCoV-19/USA/WA-S2006/2020\|EPI_ISL_497434\|2020-06-0 | .................... | .................... | ..................... | .................... | .................... | .................... |
| F | 4408 | hCoV-19/USA/WA-S2007/2020\|EPI_ISL_497435\|2020-06-0 | .................... | .................... | ..................... | .................... | .................... | .................... |
| F | 4409 | hCoV-19/USA/WA-S2008/2020\|EPI_ISL_497436\|2020-06-0 | .................... | .................... | ..................... | .................... | .................... | .................... |
| F | 4410 | hCoV-19/USA/WA-S2009/2020\|EPI_ISL_497437\|2020-06-0 | .................... | .................... | ..................... | .................... | .................... | .................... |
| F | 4411 | hCoV-19/USA/WA-S2010/2020\|EPI_ISL_497438\|2020-06-0 | .................... | .................... | ..................... | .................... | .................... | .................... |
| F | 4412 | hCoV-19/USA/WA-S2012/2020\|EPI_ISL_497440\|2020-06-0 | .................... | .................... | ..................... | .................... | .................... | .................... |
| F | 4413 | hCoV-19/USA/WA-S2013/2020\|EPI_ISL_497441\|2020-06-0 | .................... | .................... | ..................... | .................... | .................... | .................... |
| F | 4414 | hCoV-19/USA/WA-S2014/2020\|EPI_ISL_497442\|2020-06-0 | .................... | .................... | ..................... | .................... | .................... | .................... |
| F | 4415 | hCoV-19/USA/WA-S2015/2020\|EPI_ISL_497443\|2020-06-0 | .................... | .................... | ..................... | .................... | .................... | .................... |
| F | 4416 | hCoV-19/USA/WA-S2023/2020\|EPI_ISL_497451\|2020-06-0 | .................... | .................... | ..................... | .................... | .................... | .................... |
| F | 4417 | hCoV-19/USA/WA-S2038/2020\|EPI_ISL_497466\|2020-06-0 | .................... | .................... | ..................... | .................... | .................... | .................... |
| F | 4418 | hCoV-19/USA/WA-S2039/2020\|EPI_ISL_497467\|2020-06-0 | .................... | .................... | ..................... | .................... | .................... | .................... |
| F | 4419 | hCoV-19/USA/WA-S2040/2020\|EPI_ISL_497468\|2020-06-0 | .................... | .................... | ..................... | .................... | .................... | .................... |
| F | 4420 | hCoV-19/USA/WA-S2041/2020\|EPI_ISL_497469\|2020-06-0 | .................... | .................... | ..................... | .................... | .................... | .................... |
| F | 4421 | hCoV-19/USA/WA-S2042/2020\|EPI_ISL_497470\|2020-06-0 | .................... | .................... | ..................... | .................... | .................... | .................... |
| F | 4422 | hCoV-19/USA/WA-S2043/2020\|EPI_ISL_497471\|2020-06-0 | .................... | .................... | ..................... | .................... | .................... | .................... |
| F | 4423 | hCoV-19/USA/WA-S2044/2020\|EPI_ISL_497472\|2020-06-0 | .................... | .................... | ..................... | .................... | .................... | .................... |
| F | 4424 | hCoV-19/USA/WA-S2045/2020\|EPI_ISL_497473\|2020-06-0 | .................... | .................... | ..................... | .................... | .................... | .................... |
| F | 4425 | hCoV-19/USA/WA-S2046/2020\|EPI_ISL_497474\|2020-06-0 | .................... | .................... | ..................... | .................... | .................... | .................... |
| F | 4426 | hCoV-19/USA/WA-S2047/2020\|EPI_ISL_497475\|2020-06-0 | .................... | .................... | ..................... | .................... | .................... | .................... |

F 4427 hCoV-19/USA/WA-S2048/2020|EPI_ISL_497476|2020-06-0 ···················· ···················· ···················· ···················· ···················· ····················
F 4428 hCoV-19/USA/WA-S2049/2020|EPI_ISL_497477|2020-06-0 ···················· ···················· ···················· ···················· ···················· ····················
F 4429 hCoV-19/USA/WA-S2050/2020|EPI_ISL_497478|2020-06-0 ···················· ···················· ···················· ···················· ···················· ····················
F 4430 hCoV-19/USA/WA-S2051/2020|EPI_ISL_497479|2020-06-0 ···················· ···················· ···················· ···················· ···················· ····················
F 4431 hCoV-19/USA/WA-S2052/2020|EPI_ISL_497480|2020-06-0 ···················· ···················· ···················· ···················· ···················· ····················
F 4432 hCoV-19/USA/WA-S2053/2020|EPI_ISL_497481|2020-06-0 ···················· ···················· ···················· ···················· ···················· ····················
F 4433 hCoV-19/USA/WA-S2054/2020|EPI_ISL_497482|2020-06-0 ···················· ···················· ···················· ···················· ···················· ····················
F 4434 hCoV-19/USA/WA-S2055/2020|EPI_ISL_497483|2020-06-0 ···················· ···················· ···················· ···················· ···················· ····················
F 4435 hCoV-19/USA/WA-S2057/2020|EPI_ISL_497485|2020-06-0 ···················· ···················· ···················· ···················· ···················· ····················
F 4436 hCoV-19/USA/WA-S2058/2020|EPI_ISL_497486|2020-06-0 ···················· ···················· ···················· ···················· ···················· ····················
F 4437 hCoV-19/USA/WA-S2059/2020|EPI_ISL_497487|2020-06-0 ···················· ···················· ···················· ···················· ···················· ····················
F 4438 hCoV-19/USA/NC-QDX-252/2020|EPI_ISL_498731|2020-04 ···················· ···················· ···················· ···················· ···················· ····················
F 4439 hCoV-19/USA/NC-QDX-253/2020|EPI_ISL_498732|2020-04 ···················· ···················· ···················· ···················· ···················· ····················
F 4440 hCoV-19/USA/NC-QDX-254/2020|EPI_ISL_498733|2020-04 ···················· ···················· ···················· ···················· ···················· ····················
F 4441 hCoV-19/USA/NM-QDX-263/2020|EPI_ISL_498734|2020-04 ···················· ···················· ···················· ···················· ···················· ····················
F 4442 hCoV-19/USA/NM-QDX-264/2020|EPI_ISL_498735|2020-04 ···················· ···················· ···················· ···················· ···················· ····················
F 4443 hCoV-19/USA/NV-QDX-217/2020|EPI_ISL_498736|2020-04 ···················· ···················· ···················· ···················· ···················· ····················
F 4444 hCoV-19/USA/TN-QDX-246/2020|EPI_ISL_498737|2020-04 ···················· ···················· ···················· ···················· ···················· ····················
F 4445 hCoV-19/USA/TX-QDX-218/2020|EPI_ISL_498738|2020-04 ···················· ···················· ···················· ···················· ···················· ····················
F 4446 hCoV-19/USA/TX-QDX-220/2020|EPI_ISL_498739|2020-04 ···················· ···················· ···················· ···················· ···················· ····················
F 4447 hCoV-19/USA/TX-QDX-250/2020|EPI_ISL_498740|2020-04 ···················· ···················· ···················· ···················· ···················· ····················
F 4448 hCoV-19/USA/TX-QDX-255/2020|EPI_ISL_498741|2020-04 ···················· ···················· ···················· ···················· ···················· ····················
F 4449 hCoV-19/USA/TX-QDX-261/2020|EPI_ISL_498742|2020-05 ···················· ···················· ···················· ···················· ···················· ····················
F 4450 hCoV-19/USA/TX-QDX-262/2020|EPI_ISL_498743|2020-04 ···················· ···················· ···················· ···················· ···················· ····················
F 4451 hCoV-19/USA/WA-QDX-235/2020|EPI_ISL_498744|2020-04 ···················· ···················· ···················· ···················· ···················· ····················
F 4452 hCoV-19/USA/WA-QDX-236/2020|EPI_ISL_498745|2020-04 ···················· ···················· ···················· ···················· ···················· ····················
F 4453 hCoV-19/USA/WA-QDX-237/2020|EPI_ISL_498746|2020-04 ···················· ···················· ···················· ···················· ···················· ····················
F 4454 hCoV-19/USA/WA-QDX-238/2020|EPI_ISL_498747|2020-04 ···················· ···················· ···················· ···················· ···················· ····················
F 4455 hCoV-19/Australia/NSW638/2020|EPI_ISL_498748|2020- ···················· ···················· ···················· ···················· ···················· ····················
F 4456 hCoV-19/Australia/NSW639/2020|EPI_ISL_498749|2020- ···················· ···················· ···················· ···················· ···················· ····················
F 4457 hCoV-19/Australia/NSW640/2020|EPI_ISL_498750|2020- ···················· ···················· ···················· ···················· ···················· ····················
F 4458 hCoV-19/Australia/NSW641/2020|EPI_ISL_498751|2020- ···················· ···················· ···················· ···················· ···················· ····················
F 4459 hCoV-19/Australia/NSW645/2020|EPI_ISL_498764|2020- ···················· ···················· ···················· ···················· ···················· ····················
F 4460 hCoV-19/Australia/NSW646/2020|EPI_ISL_498765|2020- ···················· ···················· ···················· ···················· ···················· ····················
F 4461 hCoV-19/Australia/NSW647/2020|EPI_ISL_498766|2020- ···················· ···················· ···················· ···················· ···················· ····················
F 4462 hCoV-19/Australia/NSW648/2020|EPI_ISL_498767|2020- ···················· ···················· ···················· ···················· ···················· ····················
F 4463 hCoV-19/Australia/NSW649/2020|EPI_ISL_498768|2020- ···················· ···················· ···················· ···················· ···················· ····················
F 4464 hCoV-19/Australia/NSW650/2020|EPI_ISL_498769|2020- ···················· ···················· ···················· ···················· ···················· ····················
F 4465 hCoV-19/Australia/NSW651/2020|EPI_ISL_498770|2020- ···················· ···················· ···················· ···················· ···················· ····················
F 4466 hCoV-19/Australia/NSW652/2020|EPI_ISL_498771|2020- ···················· ···················· ···················· ···················· ···················· ····················
F 4467 hCoV-19/Australia/NSW653/2020|EPI_ISL_498772|2020- ···················· ···················· ···················· ···················· ···················· ····················
F 4468 hCoV-19/Australia/NSW654/2020|EPI_ISL_498773|2020- ···················· ···················· ···················· ···················· ···················· ····················
F 4469 hCoV-19/Australia/NSW655/2020|EPI_ISL_498774|2020- ···················· ···················· ···················· ···················· ···················· ····················
F 4470 hCoV-19/Bangladesh/BCSIR-NILMRC-338/2020|EPI_ISL_4 ···················· ···················· ···················· ···················· ···················· ····················
F 4471 hCoV-19/Malaysia/3097/2020|EPI_ISL_501200|2020-05- ···················· ···················· ···················· ···················· ···················· ····················
F 4472 hCoV-19/Malaysia/3133/2020|EPI_ISL_501201|2020-03- ···················· ···················· ···················· ···················· ···················· ····················
F 4473 hCoV-19/Malaysia/3145/2020|EPI_ISL_501202|2020-03- ···················· ···················· ···················· ···················· ···················· ····················
F 4474 hCoV-19/Malaysia/3321/2020|EPI_ISL_501203|2020-03- ···················· ···················· ···················· ···················· ···················· ····················
F 4475 hCoV-19/Malaysia/3479/2020|EPI_ISL_501204|2020-03- ···················· ···················· ···················· ···················· ···················· ····················
F 4476 hCoV-19/Malaysia/3499/2020|EPI_ISL_501205|2020-03- ···················· ···················· ···················· ···················· ···················· ····················
F 4477 hCoV-19/Malaysia/3605/2020|EPI_ISL_501206|2020-03- ···················· ·············A······ ···················· ···················· ···················· ····················
F 4478 hCoV-19/Malaysia/3611/2020|EPI_ISL_501207|2020-03- ···················· ···················· ···················· ···················· ···················· ····················
F 4479 hCoV-19/Malaysia/3703/2020|EPI_ISL_501208|2020-03- ···················· ···················· ···················· ···················· ···················· ····················
F 4480 hCoV-19/Malaysia/3706/2020|EPI_ISL_501209|2020-03- ···················· ···················· ···················· ···················· ···················· ····················
F 4481 hCoV-19/Malaysia/3998/2020|EPI_ISL_501210|2020-04- ···················· ···················· ···················· A··················· ···················· ····················
F 4482 hCoV-19/Malaysia/5056/2020|EPI_ISL_501211|2020-04- ···················· ···················· ···················· ···················· ···················· ····················
F 4483 hCoV-19/Malaysia/5425/2020|EPI_ISL_501212|2020-04- ···················· ···················· ···················· ···················· ···················· ····················
F 4484 hCoV-19/Malaysia/5815/2020|EPI_ISL_501213|2020-04- ···················· ···················· ···················· ···················· ···················· ····················
F 4485 hCoV-19/Malaysia/5822/2020|EPI_ISL_501214|2020-04- ···················· ···················· ···················· ···················· ···················· ····················
F 4486 hCoV-19/Malaysia/5906/2020|EPI_ISL_501215|2020-04- ···················· ···················· ···················· ···················· ···················· ····················
F 4487 hCoV-19/Malaysia/6088/2020|EPI_ISL_501216|2020-04- ···················· ···················· ···················· ···················· ···················· ····················
F 4488 hCoV-19/Malaysia/6149/2020|EPI_ISL_501217|2020-04- ···················· ···················· ···················· ···················· ···················· ····················
F 4489 hCoV-19/Malaysia/6216/2020|EPI_ISL_501218|2020-04- ···················· ···················· ···················· ···················· ···················· ····················
F 4490 hCoV-19/Malaysia/6306/2020|EPI_ISL_501219|2020-04- ···················· ···················· ···················· ···················· ···················· ····················
F 4491 hCoV-19/Malaysia/6359/2020|EPI_ISL_501220|2020-02- ···················· ···················· ···················· ···················· ···················· ····················
F 4492 hCoV-19/Malaysia/7618/2020|EPI_ISL_501221|2020-03- ···················· ···················· ···················· ···················· ···················· ····················
F 4493 hCoV-19/Malaysia/8451/2020|EPI_ISL_501222|2020-04- ···················· ···················· ···················· ···················· ···················· ····················
F 4494 hCoV-19/Malaysia/8454/2020|EPI_ISL_501223|2020-03- ···················· ···················· ···················· ···················· ···················· ····················
F 4495 hCoV-19/Malaysia/8816/2020|EPI_ISL_501224|2020-03- ···················· ···················· ···················· ···················· ···················· ····················
F 4496 hCoV-19/Malaysia/9136/2020|EPI_ISL_501225|2020-03- ···················· ···················· ···················· ···················· ···················· ····················
F 4497 hCoV-19/Malaysia/9593/2020|EPI_ISL_501226|2020-03- ···················· ···················· ···················· ···················· ···················· ····················
F 4498 hCoV-19/Malaysia/9857/2020|EPI_ISL_501227|2020-03- ···················· ···················· ···················· ···················· ···················· ····················
F 4499 hCoV-19/Malaysia/9886/2020|EPI_ISL_501228|2020-03- ···················· ···················· ···················· ···················· ···················· ····················
F 4500 hCoV-19/Greece/16034_HPI/2020|EPI_ISL_501230|2020- ···················· ···················· ···················· ···················· ···················· ····················
F 4501 hCoV-19/Greece/19553_HPI/2020|EPI_ISL_501233|2020- ···················· ···················· ···················· ···················· ···················· ····················
F 4502 hCoV-19/Greece/26072_HPI/2020|EPI_ISL_501236|2020- ···················· ···················· ···················· ···················· ···················· ····················
F 4503 hCoV-19/SaudiArabia/KAUST-MADINAH898/2020|EPI_ISL_ ···················· ···················· ···················· ···················· ···················· ····················
F 4504 hCoV-19/SaudiArabia/KAUST-MADINAH937/2020|EPI_ISL_ ···················· ···················· ···················· ···················· ···················· ····················
F 4505 hCoV-19/SaudiArabia/KAUST-MADINAH938/2020|EPI_ISL_ ···················· ·A·················· ···················· ···················· ···················· ··A·················
F 4506 hCoV-19/SaudiArabia/KAUST-MADINAH941/2020|EPI_ISL_ ···················· ···················· ···················· ···················· ···················· ····················

| | | | | | | | |
|---|---|---|---|---|---|---|---|
| F | 4507 hCoV-19/SaudiArabia/KAUST-MADINAH947/2020\|EPI_ISL_ | .................... | .................... | .................... | .................... | .................... | .................... |
| F | 4508 hCoV-19/SaudiArabia/KAUST-MADINAH955/2020\|EPI_ISL_ | .................... | .................... | .................... | .................... | .................... | .................... |
| F | 4509 hCoV-19/SaudiArabia/KAUST-MADINAH956/2020\|EPI_ISL_ | .................... | .................... | .................... | .................... | .................... | .................... |
| F | 4510 hCoV-19/SaudiArabia/KAUST-MADINAH957/2020\|EPI_ISL_ | .................... | .A.................. | .................... | .................... | .................... | ..A................. |
| F | 4511 hCoV-19/SaudiArabia/KAUST-MADINAH958/2020\|EPI_ISL_ | .................... | .A.................. | .................... | .................... | .................... | ..A................. |
| F | 4512 hCoV-19/SaudiArabia/KAUST-MADINAH959/2020\|EPI_ISL_ | .................... | .................... | .................... | .................... | .................... | .................... |
| F | 4513 hCoV-19/Saudi Arabia/KAUST-MADINAH960/2020\|EPI_ISL | .................... | .................... | .................... | .................... | .................... | .................... |
| F | 4514 hCoV-19/SaudiArabia/KAUST-MADINAH961/2020\|EPI_ISL_ | .................... | .................... | .................... | .................... | .................... | .................... |
| F | 4515 hCoV-19/SaudiArabia/KAUST-MADINAH967/2020\|EPI_ISL_ | .................... | .................... | .................... | .................... | .................... | .................... |
| F | 4516 hCoV-19/SaudiArabia/KAUST-MAKKAH290/2020\|EPI_ISL_5 | .................... | .A.................. | .................... | .................... | .................... | ..A................. |
| F | 4517 hCoV-19/SaudiArabia/KAUST-MAKKAH291/2020\|EPI_ISL_5 | .................... | .A.................. | .................... | .................... | .................... | ..A................. |
| F | 4518 hCoV-19/SaudiArabia/KAUST-MAKKAH293/2020\|EPI_ISL_5 | .................... | .................... | .................... | .................... | .................... | .................... |
| F | 4519 hCoV-19/SaudiArabia/KAUST-MAKKAH294/2020\|EPI_ISL_5 | .................... | .................... | .................... | .................... | .................... | .................... |
| F | 4520 hCoV-19/England/PORT-2D3DB2/2020\|EPI_ISL_514505\|20 | .................... | .................... | .................... | .................... | .................... | .................... |
| F | 4521 hCoV-19/England/PORT-2D3DC1/2020\|EPI_ISL_514506\|20 | .................... | .................... | .................... | .................... | .................... | .................... |
| F | 4522 hCoV-19/England/PORT-2D3E46/2020\|EPI_ISL_514508\|20 | .................... | .................... | .................... | .................... | .................... | .................... |
| F | 4523 hCoV-19/England/PORT-2D3E64/2020\|EPI_ISL_514510\|20 | .................... | .................... | .................... | .................... | .................... | .................... |
| F | 4524 hCoV-19/Scotland/CVR3962/2020\|EPI_ISL_514514\|2020- | .................... | .................... | .................... | .................... | .................... | .................... |
| F | 4525 hCoV-19/Scotland/CVR3963/2020\|EPI_ISL_514515\|2020- | .................... | .................... | .................... | .................... | .................... | .................... |
| F | 4526 hCoV-19/Scotland/EDB7075/2020\|EPI_ISL_514516\|2020- | .................... | .................... | .................... | .................... | .................... | .................... |
| F | 4527 hCoV-19/Scotland/EDB7076/2020\|EPI_ISL_514517\|2020- | .................... | .................... | .................... | .................... | .................... | .................... |
| F | 4528 hCoV-19/Scotland/EDB7079/2020\|EPI_ISL_514518\|2020- | .................... | .................... | .................... | .................... | .................... | .................... |
| F | 4529 hCoV-19/Scotland/EDB7080/2020\|EPI_ISL_514519\|2020- | .................... | .................... | .................... | .................... | .................... | .................... |
| F | 4530 hCoV-19/Scotland/EDB7081/2020\|EPI_ISL_514520\|2020- | .................... | .................... | .................... | .................... | .................... | .................... |
| F | 4531 hCoV-19/Scotland/EDB7084/2020\|EPI_ISL_514521\|2020- | .................... | .................... | .................... | .................... | .................... | .................... |
| F | 4532 hCoV-19/Scotland/EDB7086/2020\|EPI_ISL_514522\|2020- | .................... | .................... | .................... | .................... | .................... | .................... |
| F | 4533 hCoV-19/Scotland/EDB7088/2020\|EPI_ISL_514523\|2020- | .................... | .................... | .................... | .................... | .................... | .................... |
| F | 4534 hCoV-19/Scotland/EDB7092/2020\|EPI_ISL_514524\|2020- | .................... | .................... | .................... | .................... | .................... | .................... |
| F | 4535 hCoV-19/Scotland/EDB7094/2020\|EPI_ISL_514526\|2020- | .................... | .................... | .................... | .................... | .................... | .................... |
| F | 4536 hCoV-19/Scotland/EDB7101/2020\|EPI_ISL_514527\|2020- | .................... | .................... | .................... | .................... | .................... | .................... |
| F | 4537 hCoV-19/Scotland/EDB7102/2020\|EPI_ISL_514528\|2020- | .................... | .................... | .................... | .................... | .................... | .................... |
| F | 4538 hCoV-19/Scotland/EDB7103/2020\|EPI_ISL_514529\|2020- | .................... | .................... | .................... | .................... | .................... | .................... |
| F | 4539 hCoV-19/Scotland/EDB7104/2020\|EPI_ISL_514530\|2020- | .................... | .................... | .................... | .................... | .................... | .................... |
| F | 4540 hCoV-19/Scotland/EDB7105/2020\|EPI_ISL_514531\|2020- | .................... | .................... | .................... | .................... | .................... | .................... |
| F | 4541 hCoV-19/Scotland/EDB7109/2020\|EPI_ISL_514533\|2020- | .................... | .................... | .................... | .................... | .................... | .................... |
| F | 4542 hCoV-19/Scotland/EDB7110/2020\|EPI_ISL_514534\|2020- | .................... | .................... | .................... | .................... | .................... | .................... |
| F | 4543 hCoV-19/Scotland/EDB7111/2020\|EPI_ISL_514535\|2020- | .................... | .................... | .................... | .................... | .................... | .................... |
| F | 4544 hCoV-19/Scotland/EDB7116/2020\|EPI_ISL_514536\|2020- | .................... | .................... | .................... | .................... | .................... | .................... |
| F | 4545 hCoV-19/Scotland/EDB7118/2020\|EPI_ISL_514537\|2020- | .................... | .................... | .................... | .................... | .................... | .................... |
| F | 4546 hCoV-19/Scotland/EDB7121/2020\|EPI_ISL_514539\|2020- | .................... | .................... | .................... | .................... | .................... | .................... |
| F | 4547 hCoV-19/Scotland/EDB7123/2020\|EPI_ISL_514540\|2020- | .................... | .................... | .................... | .................... | .................... | .................... |
| F | 4548 hCoV-19/Scotland/EDB7124/2020\|EPI_ISL_514541\|2020- | .................... | .................... | .................... | .................... | .................... | .................... |
| F | 4549 hCoV-19/Scotland/EDB7126/2020\|EPI_ISL_514542\|2020- | .................... | .................... | .................... | .................... | .................... | .................... |
| F | 4550 hCoV-19/Scotland/EDB7127/2020\|EPI_ISL_514543\|2020- | .................... | .................... | .................... | .................... | .................... | .................... |
| F | 4551 hCoV-19/Scotland/EDB7132/2020\|EPI_ISL_514544\|2020- | .................... | .................... | .................... | .................... | .................... | .................... |
| F | 4552 hCoV-19/Scotland/EDB7133/2020\|EPI_ISL_514545\|2020- | .................... | .................... | .................... | .................... | .................... | .................... |
| F | 4553 hCoV-19/Scotland/EDB7135/2020\|EPI_ISL_514546\|2020- | .................... | .................... | .................... | .................... | .................... | .................... |
| F | 4554 hCoV-19/Scotland/EDB7136/2020\|EPI_ISL_514547\|2020- | .................... | .................... | .................... | .................... | .................... | .................... |
| F | 4555 hCoV-19/Scotland/EDB7139/2020\|EPI_ISL_514548\|2020- | .................... | .................... | .................... | .................... | .................... | .................... |
| F | 4556 hCoV-19/South Africa/KRISP-K001340/2020\|EPI_ISL_51 | .................... | .................... | .................... | .................... | .................... | .................... |
| F | 4557 hCoV-19/South Africa/KRISP-K001341/2020\|EPI_ISL_51 | .................... | .................... | .................... | .................... | .................... | .................... |
| F | 4558 hCoV-19/South Africa/KRISP-K001342/2020\|EPI_ISL_51 | .................... | .................... | .................... | .................... | .................... | .................... |
| F | 4559 hCoV-19/South Africa/KRISP-K001343/2020\|EPI_ISL_51 | .................... | .................... | .................... | .................... | .................... | .................... |
| F | 4560 hCoV-19/South Africa/KRISP-K001344/2020\|EPI_ISL_51 | .................... | .................... | .................... | .................... | .................... | .................... |
| F | 4561 hCoV-19/South Africa/KRISP-K001345/2020\|EPI_ISL_51 | .................... | .................... | .................... | .................... | .................... | .................... |
| F | 4562 hCoV-19/South Africa/KRISP-K001346/2020\|EPI_ISL_51 | .................... | .................... | .................... | .................... | .................... | .................... |
| F | 4563 hCoV-19/South Africa/KRISP-K001347/2020\|EPI_ISL_51 | .................... | .................... | .................... | .................... | .................... | .................... |
| F | 4564 hCoV-19/South Africa/KRISP-K001348/2020\|EPI_ISL_51 | .................... | .................... | .................... | .................... | .................... | .................... |
| F | 4565 hCoV-19/South Africa/KRISP-K001350/2020\|EPI_ISL_51 | .................... | .................... | .................... | .................... | .................... | .................... |
| F | 4566 hCoV-19/South Africa/KRISP-K001351/2020\|EPI_ISL_51 | .................... | .................... | .................... | .................... | .................... | .................... |
| F | 4567 hCoV-19/South Africa/KRISP-K001352/2020\|EPI_ISL_51 | .................... | .................... | .................... | .................... | .................... | .................... |
| F | 4568 hCoV-19/South Africa/KRISP-K001353/2020\|EPI_ISL_51 | .................... | .................... | .................... | .................... | .................... | .................... |
| F | 4569 hCoV-19/South Africa/KRISP-K001354/2020\|EPI_ISL_51 | .................... | .................... | .................... | .................... | .................... | .................... |
| F | 4570 hCoV-19/South Africa/KRISP-K001355/2020\|EPI_ISL_51 | .................... | .................... | .................... | .................... | .................... | .................... |
| F | 4571 hCoV-19/South Africa/KRISP-K001356/2020\|EPI_ISL_51 | .................... | .................... | .................... | .................... | .................... | .................... |
| F | 4572 hCoV-19/South Africa/KRISP-K001357/2020\|EPI_ISL_51 | .................... | .................... | .................... | .................... | .................... | .................... |
| F | 4573 hCoV-19/South Africa/KRISP-K001358/2020\|EPI_ISL_51 | .................... | .................... | .................... | .................... | .................... | .................... |
| F | 4574 hCoV-19/South Africa/KRISP-K001359/2020\|EPI_ISL_51 | .................... | .................... | .................... | .................... | .................... | .................... |
| F | 4575 hCoV-19/South Africa/KRISP-K001360/2020\|EPI_ISL_51 | .................... | .................... | .................... | .................... | .................... | .................... |
| F | 4576 hCoV-19/South Africa/KRISP-K001361/2020\|EPI_ISL_51 | .................... | .................... | .................... | .................... | .................... | .................... |
| F | 4577 hCoV-19/South Africa/KRISP-K001362/2020\|EPI_ISL_51 | .................... | .................... | .................... | .................... | .................... | .................... |
| F | 4578 hCoV-19/South Africa/KRISP-K001363/2020\|EPI_ISL_51 | .................... | .................... | .................... | .................... | .................... | .................... |
| F | 4579 hCoV-19/South Africa/KRISP-K001364/2020\|EPI_ISL_51 | .................... | .................... | .................... | .................... | .................... | .................... |
| F | 4580 hCoV-19/South Africa/KRISP-K001365/2020\|EPI_ISL_51 | .................... | .................... | .................... | .................... | .................... | .................... |
| F | 4581 hCoV-19/South Africa/KRISP-K001366/2020\|EPI_ISL_51 | .................... | .................... | .................... | .................... | .................... | .................... |
| F | 4582 hCoV-19/South Africa/KRISP-K001367/2020\|EPI_ISL_51 | .................... | .................... | .................... | .................... | .................... | .................... |
| F | 4583 hCoV-19/South Africa/KRISP-K001368/2020\|EPI_ISL_51 | .................... | .................... | .................... | .................... | .................... | .................... |
| F | 4584 hCoV-19/South Africa/KRISP-K001369/2020\|EPI_ISL_51 | .................... | .................... | .................... | .................... | .................... | .................... |
| F | 4585 hCoV-19/South Africa/KRISP-K001370/2020\|EPI_ISL_51 | .................... | .................... | .................... | .................... | .................... | .................... |
| F | 4586 hCoV-19/South Africa/KRISP-K001371/2020\|EPI_ISL_51 | .................... | .................... | .................... | .................... | .................... | .................... |

| | | | | | | | | |
|---|---|---|---|---|---|---|---|---|
| F | 4587 | hCoV-19/South Africa/KRISP-K001372/2020\|EPI_ISL_51 | ···················· | ···················· | ···················· | ···················· | ···················· | ···················· |
| F | 4588 | hCoV-19/South Africa/KRISP-K001373/2020\|EPI_ISL_51 | ···················· | ···················· | ···················· | ···················· | ···················· | ···················· |
| F | 4589 | hCoV-19/South Africa/KRISP-K001374/2020\|EPI_ISL_51 | ···················· | ···················· | ···················· | ···················· | ···················· | ···················· |
| F | 4590 | hCoV-19/South Africa/KRISP-K001375/2020\|EPI_ISL_51 | ···················· | ···················· | ···················· | ···················· | ···················· | ···················· |
| F | 4591 | hCoV-19/South Africa/KRISP-K001376/2020\|EPI_ISL_51 | ···················· | ···················· | ···················· | ···················· | ···················· | ···················· |
| F | 4592 | hCoV-19/South Africa/KRISP-K001377/2020\|EPI_ISL_51 | ···················· | ···················· | ···················· | ···················· | ···················· | ···················· |
| F | 4593 | hCoV-19/South Africa/KRISP-K001378/2020\|EPI_ISL_51 | ···················· | ···················· | ···················· | ···················· | ···················· | ···················· |
| F | 4594 | hCoV-19/South Africa/KRISP-K001379/2020\|EPI_ISL_51 | ···················· | ···················· | ···················· | ···················· | ···················· | ···················· |
| F | 4595 | hCoV-19/South Africa/KRISP-K001380/2020\|EPI_ISL_51 | ···················· | ···················· | ···················· | ···················· | ···················· | ···················· |
| F | 4596 | hCoV-19/South Africa/KRISP-K001381/2020\|EPI_ISL_51 | ···················· | ···················· | ···················· | ···················· | ···················· | ···················· |
| F | 4597 | hCoV-19/South Africa/KRISP-K001383/2020\|EPI_ISL_51 | ···················· | ···················· | ···················· | ···················· | ···················· | ···················· |
| F | 4598 | hCoV-19/South Africa/KRISP-K001384/2020\|EPI_ISL_51 | ········T··········· | ···················· | ···················· | ···················· | ···················· | ···················· |
| F | 4599 | hCoV-19/South Africa/KRISP-K001385/2020\|EPI_ISL_51 | ···················· | ···················· | ···················· | ···················· | ···················· | ···················· |
| F | 4600 | hCoV-19/South Africa/KRISP-K001386/2020\|EPI_ISL_51 | ···················· | ···················· | ···················· | ···················· | ···················· | ···················· |
| F | 4601 | hCoV-19/South Africa/KRISP-K001387/2020\|EPI_ISL_51 | ···················· | ···················· | ···················· | ···················· | ···················· | ···················· |
| F | 4602 | hCoV-19/South Africa/KRISP-K001388/2020\|EPI_ISL_51 | ···················· | ···················· | ···················· | ···················· | ···················· | ···················· |
| F | 4603 | hCoV-19/South Africa/KRISP-K001389/2020\|EPI_ISL_51 | ···················· | ···················· | ···················· | ···················· | ···················· | ···················· |
| F | 4604 | hCoV-19/South Africa/KRISP-K001390/2020\|EPI_ISL_51 | ···················· | ···················· | ···················· | ···················· | ···················· | ···················· |
| F | 4605 | hCoV-19/South Africa/KRISP-K001391/2020\|EPI_ISL_51 | ···················· | ···················· | ···················· | ···················· | ···················· | ···················· |
| F | 4606 | hCoV-19/South Africa/KRISP-K001392/2020\|EPI_ISL_51 | ········T··········· | ···················· | ···················· | ···················· | ···················· | ···················· |
| F | 4607 | hCoV-19/South Africa/KRISP-K001393/2020\|EPI_ISL_51 | ···················· | ···················· | ···················· | ···················· | ···················· | ···················· |
| F | 4608 | hCoV-19/South Africa/KRISP-K001394/2020\|EPI_ISL_51 | ···················· | ···················· | ···················· | ···················· | ···················· | ···················· |
| F | 4609 | hCoV-19/USA/WA-S1674/2020\|EPI_ISL_496100\|2020-05-2 | ···················· | ···················· | ···················· | ···················· | ···················· | ···················· |
| F | 4610 | hCoV-19/USA/WA-S1675/2020\|EPI_ISL_496101\|2020-05-2 | ···················· | ···················· | ···················· | ···················· | ···················· | ···················· |
| F | 4611 | hCoV-19/USA/WA-S1684/2020\|EPI_ISL_496110\|2020-05-3 | ···················· | ···················· | ···················· | ···················· | ···················· | ···················· |
| F | 4612 | hCoV-19/USA/WA-S1994/2020\|EPI_ISL_497422\|2020-06-0 | ···················· | ···················· | ···················· | ···················· | ···················· | ···················· |
| F | 4613 | hCoV-19/USA/WA-S1995/2020\|EPI_ISL_497423\|2020-06-0 | ···················· | ···················· | ···················· | ···················· | ···················· | ···················· |
| F | 4614 | hCoV-19/USA/WA-S1996/2020\|EPI_ISL_497424\|2020-06-0 | ···················· | ···················· | ···················· | ···················· | ···················· | ···················· |
| F | 4615 | hCoV-19/USA/WA-S1997/2020\|EPI_ISL_497425\|2020-06-0 | ···················· | ···················· | ···················· | ···················· | ···················· | ···················· |
| F | 4616 | hCoV-19/USA/WA-S1998/2020\|EPI_ISL_497426\|2020-06-0 | ···················· | ···················· | ···················· | ···················· | ···················· | ···················· |
| F | 4617 | hCoV-19/USA/WA-S1999/2020\|EPI_ISL_497427\|2020-06-0 | ···················· | ···················· | ···················· | ···················· | ···················· | ···················· |
| F | 4618 | hCoV-19/USA/WA-S2000/2020\|EPI_ISL_497428\|2020-06-0 | ···················· | ···················· | ···················· | ···················· | ···················· | ···················· |
| F | 4619 | hCoV-19/USA/WA-S2001/2020\|EPI_ISL_497429\|2020-06-0 | ···················· | ···················· | ···················· | ···················· | ···················· | ···················· |
| F | 4620 | hCoV-19/USA/WA-S2002/2020\|EPI_ISL_497430\|2020-06-0 | ···················· | ···················· | ···················· | ···················· | ···················· | ···················· |
| F | 4621 | hCoV-19/USA/WA-S2003/2020\|EPI_ISL_497431\|2020-06-0 | ···················· | ···················· | ···················· | ···················· | ···················· | ···················· |
| F | 4622 | hCoV-19/USA/WA-S2004/2020\|EPI_ISL_497432\|2020-06-0 | ···················· | ···················· | ···················· | ···················· | ···················· | ···················· |
| F | 4623 | hCoV-19/USA/CA-QDX-265/2020\|EPI_ISL_498700\|2020-05 | ···················· | ···················· | ···················· | ···················· | ···················· | ···················· |
| F | 4624 | hCoV-19/USA/CA-QDX-266/2020\|EPI_ISL_498701\|2020-05 | ···················· | ···················· | ···················· | ···················· | ···················· | ···················· |
| F | 4625 | hCoV-19/USA/CA-QDX-267/2020\|EPI_ISL_498702\|2020-05 | ···················· | ···················· | ···················· | ···················· | ···················· | ···················· |
| F | 4626 | hCoV-19/USA/CA-QDX-268/2020\|EPI_ISL_498703\|2020-05 | ···················· | ···················· | ···················· | ···················· | ···················· | ···················· |
| F | 4627 | hCoV-19/USA/CA-QDX-269/2020\|EPI_ISL_498704\|2020-05 | ···················· | ···················· | ···················· | ···················· | ···················· | ···················· |
| F | 4628 | hCoV-19/USA/FL-QDX-224/2020\|EPI_ISL_498705\|2020-04 | ···················· | ···················· | ···················· | ···················· | ···················· | ···················· |
| F | 4629 | hCoV-19/USA/FL-QDX-225/2020\|EPI_ISL_498706\|2020-04 | ···················· | ···················· | ···················· | ···················· | ···················· | ···················· |
| F | 4630 | hCoV-19/USA/FL-QDX-226/2020\|EPI_ISL_498707\|2020-04 | ···················· | ···················· | ···················· | ···················· | ···················· | ···················· |
| F | 4631 | hCoV-19/USA/FL-QDX-227/2020\|EPI_ISL_498708\|2020-04 | ···················· | ···················· | ···················· | ···················· | ···················· | ···················· |
| F | 4632 | hCoV-19/USA/FL-QDX-228/2020\|EPI_ISL_498709\|2020-04 | ···················· | ···················· | ···················· | ···················· | ···················· | ···················· |
| F | 4633 | hCoV-19/USA/FL-QDX-229/2020\|EPI_ISL_498710\|2020-04 | ···················· | ···················· | ···················· | ···················· | ···················· | ···················· |
| F | 4634 | hCoV-19/USA/FL-QDX-230/2020\|EPI_ISL_498711\|2020-04 | ···················· | ···················· | ···················· | ···················· | ···················· | ···················· |
| F | 4635 | hCoV-19/USA/FL-QDX-231/2020\|EPI_ISL_498712\|2020-04 | ···················· | ···················· | ···················· | ···················· | ···················· | ···················· |
| F | 4636 | hCoV-19/USA/GA-QDX-232/2020\|EPI_ISL_498713\|2020-04 | ···················· | ···················· | ···················· | ···················· | ···················· | ···················· |
| F | 4637 | hCoV-19/USA/GA-QDX-233/2020\|EPI_ISL_498714\|2020-04 | ···················· | ···················· | ···················· | ···················· | ···················· | ···················· |
| F | 4638 | hCoV-19/USA/GA-QDX-234/2020\|EPI_ISL_498715\|2020-04 | ···················· | ···················· | ···················· | ···················· | ···················· | ···················· |
| F | 4639 | hCoV-19/USA/GA-QDX-239/2020\|EPI_ISL_498716\|2020-04 | ···················· | ····A··············· | ···················· | ···················· | ···················· | ···················· |
| F | 4640 | hCoV-19/USA/GA-QDX-240/2020\|EPI_ISL_498717\|2020-04 | ···················· | ···················· | ···················· | ···················· | ···················· | ···················· |
| F | 4641 | hCoV-19/USA/GA-QDX-241/2020\|EPI_ISL_498718\|2020-04 | ···················· | ·········A·········· | ···················· | ···················· | ···················· | ···················· |
| F | 4642 | hCoV-19/USA/GA-QDX-242/2020\|EPI_ISL_498719\|2020-04 | ···················· | ···················· | ···················· | ···················· | ···················· | ···················· |
| F | 4643 | hCoV-19/USA/GA-QDX-243/2020\|EPI_ISL_498720\|2020-04 | ···················· | ···················· | ···················· | ···················· | ···················· | ···················· |
| F | 4644 | hCoV-19/USA/IA-QDX-249/2020\|EPI_ISL_498721\|2020-04 | ···················· | ···················· | ···················· | ···················· | ···················· | ···················· |
| F | 4645 | hCoV-19/USA/IL-QDX-248/2020\|EPI_ISL_498722\|2020-04 | ···················· | ···················· | ···················· | ···················· | ···················· | ···················· |
| F | 4646 | hCoV-19/USA/IL-QDX-258/2020\|EPI_ISL_498723\|2020-04 | ···················· | ···················· | ···················· | ···················· | ···················· | ···················· |
| F | 4647 | hCoV-19/USA/IL-QDX-259/2020\|EPI_ISL_498724\|2020-04 | ···················· | ···················· | ···················· | ···················· | ···················· | ···················· |
| F | 4648 | hCoV-19/USA/IL-QDX-260/2020\|EPI_ISL_498725\|2020-04 | ···················· | ···················· | ···················· | ···················· | ···················· | ···················· |
| F | 4649 | hCoV-19/USA/IN-QDX-251/2020\|EPI_ISL_498726\|2020-04 | ···················· | ···················· | ···················· | ···················· | ···················· | ···················· |
| F | 4650 | hCoV-19/USA/LA-QDX-256/2020\|EPI_ISL_498727\|2020-04 | ···················· | ···················· | ···················· | ···················· | ···················· | ···················· |
| F | 4651 | hCoV-19/USA/MI-QDX-216/2020\|EPI_ISL_498728\|2020-03 | ···················· | ···················· | ···················· | ···················· | ···················· | ···················· |
| F | 4652 | hCoV-19/USA/MN-QDX-257/2020\|EPI_ISL_498729\|2020-04 | ···················· | ···················· | ···················· | ···················· | ···················· | ···················· |
| F | 4653 | hCoV-19/USA/MS-QDX-247/2020\|EPI_ISL_498730\|2020-04 | ···················· | ···················· | ···················· | ···················· | ···················· | ···················· |
| F | 4654 | hCoV-19/Australia/NSW642/2020\|EPI_ISL_498761\|2020- | ···················· | ···················· | ···················· | ···················· | ···················· | ···················· |
| F | 4655 | hCoV-19/Australia/NSW643/2020\|EPI_ISL_498762\|2020- | ···················· | ···················· | ···················· | ···················· | ···················· | ···················· |
| F | 4656 | hCoV-19/Australia/NSW644/2020\|EPI_ISL_498763\|2020- | ···················· | ···················· | ···················· | ···················· | ···················· | ···················· |
| F | 4657 | hCoV-19/Ireland/D-NVRL-20IRL43203/2020\|EPI_ISL_501 | ···················· | ·A·················· | ···················· | ···················· | ···················· | ···A················ |
| F | 4658 | hCoV-19/Ireland/D-NVRL-20IRL47314/2020\|EPI_ISL_501 | ···················· | ···················· | ···················· | ···················· | ···················· | ···················· |
| F | 4659 | hCoV-19/Ireland/D-NVRL-20IRL47363/2020\|EPI_ISL_501 | ···················· | ···················· | ···················· | ···················· | ···················· | ···················· |
| F | 4660 | hCoV-19/Latvia/067/2020\|EPI_ISL_501275\|2020-07-08 | ···················· | ···················· | ···················· | ···················· | ···················· | ···················· |
| F | 4661 | hCoV-19/Latvia/068/2020\|EPI_ISL_501284\|2020-07-10 | ···················· | ···················· | ···················· | ···················· | ···················· | ···················· |
| F | 4662 | hCoV-19/Latvia/069/2020\|EPI_ISL_501285\|2020-07-10 | ···················· | ···················· | ···················· | ···················· | ···················· | ···················· |
| F | 4663 | hCoV-19/Latvia/070/2020\|EPI_ISL_501286\|2020-06-05 | ···················· | ···················· | ···················· | ···················· | ···················· | ···················· |
| F | 4664 | hCoV-19/Latvia/071/2020\|EPI_ISL_501287\|2020-06-06 | ···················· | ···················· | ···················· | ···················· | ···················· | ···················· |
| F | 4665 | hCoV-19/Latvia/072/2020\|EPI_ISL_501288\|2020-07-10 | ···················· | ···················· | ···················· | ···················· | ···················· | ···················· |
| F | 4666 | hCoV-19/Latvia/073/2020\|EPI_ISL_501289\|2020-07-10 | ···················· | ···················· | ···················· | ···················· | ···················· | ···················· |

| | | | | | | | |
|---|---|---|---|---|---|---|---|
| F | 4667 hCoV-19/Russia/StPetersburg-RII17940S/2020\|EPI_ISL | .................... | .................... | ..................... | .................... | .................... | .................... |
| F | 4668 hCoV-19/Russia/StPetersburg-RII17941S/2020\|EPI_ISL | .................... | .................... | ..................... | .................... | .................... | .................... |
| F | 4669 hCoV-19/Russia/StPetersburg-RII18088S/2020\|EPI_ISL | .................... | .................... | ..................... | .................... | .................... | .................... |
| F | 4670 hCoV-19/Russia/StPetersburg-RII18089S/2020\|EPI_ISL | .................... | .................... | ..................... | .................... | .................... | .................... |
| F | 4671 hCoV-19/Russia/StPetersburg-RII18090S/2020\|EPI_ISL | .................... | .................... | ..................... | .................... | .................... | .................... |
| F | 4672 hCoV-19/Russia/StPetersburg-RII21538S/2020\|EPI_ISL | .................... | .................... | ..................... | .................... | .................... | .................... |
| F | 4673 hCoV-19/Russia/StPetersburg-RII21769S/2020\|EPI_ISL | .................... | .................... | ..................... | .................... | .................... | .................... |
| F | 4674 hCoV-19/Russia/StPetersburg-RII21797S/2020\|EPI_ISL | .................... | .................... | ..................... | .................... | .................... | .................... |
| F | 4683 hCoV-19/USA/MI-MDHHS-SC21188/2020\|EPI_ISL_507445\|2 | .................... | .................... | ..................... | .................... | .................... | .................... |
| F | 4696 hCoV-19/Germany/BW-UT-025/2020\|EPI_ISL_508700\|2020 | .................... | .................... | ..................... | .................... | .................... | .................... |
| F | 4697 hCoV-19/Germany/BW-UT-026/2020\|EPI_ISL_508701\|2020 | .................... | .................... | ..................... | .................... | .................... | .................... |
| F | 4698 hCoV-19/Germany/BW-UT-027/2020\|EPI_ISL_508702\|2020 | .................... | .................... | ..................... | .................... | .................... | .................... |
| F | 4699 hCoV-19/Germany/BW-UT-028/2020\|EPI_ISL_508703\|2020 | .................... | .................... | ..................... | .................... | .................... | .................... |
| F | 4700 hCoV-19/Germany/BW-UT-029/2020\|EPI_ISL_508704\|2020 | .................... | .................... | ..................... | .................... | .................... | .................... |
| F | 4701 hCoV-19/USA/FL-BPHL-0505/2020\|EPI_ISL_508708\|2020- | .................... | .................... | ..................... | .................... | .................... | .................... |
| F | 4702 hCoV-19/USA/FL-BPHL-0506/2020\|EPI_ISL_508709\|2020- | .................... | .................... | ..................... | .................... | .................... | .................... |
| F | 4703 hCoV-19/USA/FL-BPHL-0508/2020\|EPI_ISL_508711\|2020- | .................... | .................... | ..................... | .................... | .................... | .................... |
| F | 4704 hCoV-19/USA/FL-BPHL-0509/2020\|EPI_ISL_508712\|2020- | .................... | .................... | ..................... | .................... | .................... | .................... |
| F | 4706 hCoV-19/USA/FL-BPHL-0513/2020\|EPI_ISL_508716\|2020- | .................... | .................... | ..................... | .................... | .................... | .................... |
| F | 4707 hCoV-19/USA/FL-BPHL-0514/2020\|EPI_ISL_508717\|2020- | .................... | .................... | ..................... | .................... | .................... | .................... |
| F | 4708 hCoV-19/USA/FL-BPHL-0515/2020\|EPI_ISL_508718\|2020- | .................... | .................... | ..................... | .................... | .................... | .................... |
| F | 4709 hCoV-19/USA/FL-BPHL-0516/2020\|EPI_ISL_508719\|2020- | .................... | .................... | ..................... | .................... | .................... | .................... |
| F | 4711 hCoV-19/USA/FL-BPHL-0518/2020\|EPI_ISL_508721\|2020- | .................... | .................... | ..................... | .................... | .................... | .................... |
| F | 4712 hCoV-19/USA/FL-BPHL-0520/2020\|EPI_ISL_508723\|2020- | .................... | .................... | ..................... | .................... | .................... | .................... |
| F | 4713 hCoV-19/USA/FL-BPHL-0521/2020\|EPI_ISL_508724\|2020- | .................... | .................... | ..................... | .................... | .................... | .................... |
| F | 4714 hCoV-19/USA/FL-BPHL-0522/2020\|EPI_ISL_508725\|2020- | .................... | .................... | ..................... | .................... | .................... | .................... |
| F | 4715 hCoV-19/USA/FL-BPHL-0523/2020\|EPI_ISL_508726\|2020- | .................... | .................... | ..................... | .................... | .................... | .................... |
| F | 4716 hCoV-19/USA/FL-BPHL-0524/2020\|EPI_ISL_508727\|2020- | .................... | .................... | ..................... | .................... | .................... | .................... |
| F | 4717 hCoV-19/USA/FL-BPHL-0526/2020\|EPI_ISL_508729\|2020- | .................... | .................... | ..................... | .................... | .................... | .................... |
| F | 4718 hCoV-19/USA/FL-BPHL-0527/2020\|EPI_ISL_508730\|2020- | .................... | .................... | ..................... | .................... | .................... | .................... |
| F | 4719 hCoV-19/USA/FL-BPHL-0528/2020\|EPI_ISL_508731\|2020- | .................... | .................... | ..................... | .................... | .................... | .................... |
| F | 4720 hCoV-19/USA/FL-BPHL-0529/2020\|EPI_ISL_508732\|2020- | .................... | .................... | ..................... | .................... | .................... | .................... |
| F | 4721 hCoV-19/USA/FL-BPHL-0530/2020\|EPI_ISL_508733\|2020- | .................... | .................... | ..................... | .................... | .................... | .................... |
| F | 4722 hCoV-19/USA/FL-BPHL-0531/2020\|EPI_ISL_508734\|2020- | .................... | .............T...... | ..................... | .................... | .................... | .................... |
| F | 4723 hCoV-19/USA/FL-BPHL-0532/2020\|EPI_ISL_508735\|2020- | .................... | .................... | ..................... | .................... | .................... | .................... |
| F | 4724 hCoV-19/USA/FL-BPHL-0533/2020\|EPI_ISL_508736\|2020- | .................... | .................... | ..................... | .................... | .................... | .................... |
| F | 4725 hCoV-19/USA/FL-BPHL-0534/2020\|EPI_ISL_508737\|2020- | .................... | .................... | ..................... | .................... | .................... | .................... |
| F | 4726 hCoV-19/USA/FL-BPHL-0540/2020\|EPI_ISL_508743\|2020- | .................... | .................... | ..................... | .................... | .................... | .................... |
| F | 4727 hCoV-19/USA/FL-BPHL-0541/2020\|EPI_ISL_508744\|2020- | .................... | .................... | ..................... | .................... | .................... | .................... |
| F | 4728 hCoV-19/USA/FL-BPHL-0542/2020\|EPI_ISL_508745\|2020- | .................... | .................... | ..................... | .................... | .................... | .................... |
| F | 4729 hCoV-19/USA/FL-BPHL-0543/2020\|EPI_ISL_508746\|2020- | .................... | .................... | ..................... | .................... | .................... | .................... |
| F | 4730 hCoV-19/USA/FL-BPHL-0545/2020\|EPI_ISL_508748\|2020- | .................... | .................... | ..................... | .................... | .................... | .................... |
| F | 4731 hCoV-19/USA/FL-BPHL-0546/2020\|EPI_ISL_508749\|2020- | .................... | .................... | ..................... | .................... | .................... | .................... |
| F | 4732 hCoV-19/USA/FL-BPHL-0547/2020\|EPI_ISL_508750\|2020- | .................... | .................... | ..................... | .................... | .................... | .................... |
| F | 4733 hCoV-19/USA/FL-BPHL-0549/2020\|EPI_ISL_508752\|2020- | .................... | .................... | ..................... | .................... | .................... | .................... |
| F | 4734 hCoV-19/USA/FL-BPHL-0550/2020\|EPI_ISL_508753\|2020- | .................... | .................... | ..................... | .................... | .................... | .................... |
| F | 4735 hCoV-19/USA/FL-BPHL-0551/2020\|EPI_ISL_508754\|2020- | .................... | .................... | ..................... | .................... | .................... | .................... |
| F | 4736 hCoV-19/USA/FL-BPHL-0552/2020\|EPI_ISL_508755\|2020- | .................... | .................... | ..................... | .................... | .................... | .................... |
| F | 4737 hCoV-19/USA/FL-BPHL-0553/2020\|EPI_ISL_508756\|2020- | .................... | .................... | ..................... | .................... | .................... | .................... |
| F | 4738 hCoV-19/USA/FL-BPHL-0554/2020\|EPI_ISL_508757\|2020- | .................... | .................... | ..................... | .................... | .................... | .................... |
| F | 4739 hCoV-19/USA/FL-BPHL-0555/2020\|EPI_ISL_508758\|2020- | .................... | .................... | ..................... | .................... | .................... | .................... |
| F | 4740 hCoV-19/USA/FL-BPHL-0556/2020\|EPI_ISL_508759\|2020- | .................... | .................... | ..................... | .................... | .................... | .................... |
| F | 4741 hCoV-19/USA/FL-BPHL-0557/2020\|EPI_ISL_508760\|2020- | .................... | .................... | ..................... | .................... | .................... | .................... |
| F | 4742 hCoV-19/USA/FL-BPHL-0559/2020\|EPI_ISL_508762\|2020- | .................... | .................... | ..................... | .................... | .................... | .................... |
| F | 4743 hCoV-19/USA/FL-BPHL-0561/2020\|EPI_ISL_508765\|2020- | .................... | .................... | ..................... | .................... | .................... | .................... |
| F | 4744 hCoV-19/USA/FL-BPHL-0562/2020\|EPI_ISL_508766\|2020- | .................... | .................... | ..................... | .................... | .................... | .................... |
| F | 4745 hCoV-19/USA/FL-BPHL-0563/2020\|EPI_ISL_508767\|2020- | .................... | .................... | ..................... | .................... | .................... | .................... |
| F | 4746 hCoV-19/USA/FL-BPHL-0565/2020\|EPI_ISL_508769\|2020- | .................... | .................... | ..................... | .................... | .................... | .................... |
| F | 4747 hCoV-19/USA/FL-BPHL-0567/2020\|EPI_ISL_508771\|2020- | .................... | .................... | ..................... | .................... | .................... | .................... |
| F | 4748 hCoV-19/USA/FL-BPHL-0568/2020\|EPI_ISL_508772\|2020- | .................... | .................... | ..................... | .................... | .................... | .................... |
| F | 4749 hCoV-19/USA/FL-BPHL-0569/2020\|EPI_ISL_508773\|2020- | .................... | .................... | ..................... | .................... | .................... | .................... |
| F | 4751 hCoV-19/USA/FL-BPHL-0571/2020\|EPI_ISL_508775\|2020- | .................... | .................... | ..................... | .................... | .................... | .................... |
| F | 4753 hCoV-19/USA/FL-BPHL-0573/2020\|EPI_ISL_508777\|2020- | .................... | .................... | ..................... | .................... | .................... | .................... |
| F | 4754 hCoV-19/USA/FL-BPHL-0574/2020\|EPI_ISL_508778\|2020- | .................... | .................... | ..................... | .................... | .................... | .................... |
| F | 4756 hCoV-19/USA/FL-BPHL-0576/2020\|EPI_ISL_508780\|2020- | .................... | .................... | ..................... | .................... | .................... | .................... |
| F | 4757 hCoV-19/USA/FL-BPHL-0577/2020\|EPI_ISL_508781\|2020- | .................... | .................... | ..................... | .................... | .................... | .................... |
| F | 4758 hCoV-19/USA/FL-BPHL-0578/2020\|EPI_ISL_508782\|2020- | .................... | .................... | ..................... | .................... | .................... | .................... |
| F | 4759 hCoV-19/USA/FL-BPHL-0580/2020\|EPI_ISL_508784\|2020- | .................... | .................... | ..................... | .................... | N................... | .................... |
| F | 4760 hCoV-19/USA/FL-BPHL-0582/2020\|EPI_ISL_508786\|2020- | .................... | .................... | ..................... | .................... | .................... | .................... |
| F | 4761 hCoV-19/USA/FL-BPHL-0583/2020\|EPI_ISL_508787\|2020- | .................... | .................... | ..................... | .................... | .................... | .................... |
| F | 4762 hCoV-19/USA/FL-BPHL-0584/2020\|EPI_ISL_508788\|2020- | .................... | .................... | ..................... | .................... | .................... | .................... |
| F | 4763 hCoV-19/USA/FL-BPHL-0586/2020\|EPI_ISL_508790\|2020- | .................... | .................... | ..................... | .................... | .................... | .................... |
| F | 4764 hCoV-19/USA/FL-BPHL-0587/2020\|EPI_ISL_508791\|2020- | .................... | .................... | ..................... | .................... | .................... | .................... |
| F | 4765 hCoV-19/USA/FL-BPHL-0588/2020\|EPI_ISL_508792\|2020- | .................... | .................... | ..................... | .................... | .................... | .................... |
| F | 4766 hCoV-19/USA/FL-BPHL-0589/2020\|EPI_ISL_508793\|2020- | .................... | .........A.......... | ..................... | .................... | .................... | .................... |
| F | 4767 hCoV-19/USA/FL-BPHL-0590/2020\|EPI_ISL_508794\|2020- | .................... | .................... | ..................... | .................... | .................... | .................... |
| F | 4768 hCoV-19/USA/FL-BPHL-0591/2020\|EPI_ISL_508795\|2020- | .................... | .................... | ..................... | .................... | .................... | .................... |
| F | 4769 hCoV-19/USA/FL-BPHL-0592/2020\|EPI_ISL_508796\|2020- | .................... | .................... | ..................... | .................... | .................... | .................... |
| F | 4770 hCoV-19/USA/FL-BPHL-0593/2020\|EPI_ISL_508797\|2020- | .................... | .................... | ..................... | .................... | .................... | .................... |
| F | 4771 hCoV-19/USA/FL-BPHL-0594/2020\|EPI_ISL_508798\|2020- | .................... | .................... | ..................... | .................... | .................... | .................... |

| | | | | | | | | | |
|---|---|---|---|---|---|---|---|---|---|
| F | 4772 hCoV-19/USA/FL-BPHL-0595/2020\|EPI_ISL_508799\|2020- | ···················· | ···················· | ···················· | ···················· | ···················· | ···················· | | |
| F | 4773 hCoV-19/SaudiArabia/KAUST-MAKKAH770/2020\|EPI_ISL_5 | ···················· | ···················· | ···················· | ···················· | ···················· | ···················· | | |
| F | 4774 hCoV-19/SaudiArabia/KAUST-MAKKAH771/2020\|EPI_ISL_5 | ···················· | ·A·················· | ···················· | ···················· | ···················· | ··A················· | | |
| F | 4775 hCoV-19/SaudiArabia/KAUST-MAKKAH772/2020\|EPI_ISL_5 | ···················· | ·A·················· | ···················· | ···················· | ···················· | ··A················· | | |
| F | 4776 hCoV-19/SaudiArabia/KAUST-MAKKAH773/2020\|EPI_ISL_5 | ···················· | ·A·················· | ···················· | ···················· | ···················· | ··A················· | | |
| F | 4777 hCoV-19/SaudiArabia/KAUST-MAKKAH774/2020\|EPI_ISL_5 | ···················· | ···················· | ···················· | ···················· | ···················· | ···················· | | |
| F | 4778 hCoV-19/SaudiArabia/KAUST-MAKKAH776/2020\|EPI_ISL_5 | ···················· | ·A·················· | ···················· | ···················· | ···················· | ··A················· | | |
| F | 4779 hCoV-19/SaudiArabia/KAUST-MAKKAH777/2020\|EPI_ISL_5 | ···················· | ···················· | ···················· | ···················· | ···················· | ···················· | | |
| F | 4780 hCoV-19/SaudiArabia/KAUST-MAKKAH778/2020\|EPI_ISL_5 | ···················· | ·A·················· | ···················· | ···················· | ···················· | ··A················· | | |
| F | 4781 hCoV-19/Saudi Arabia/KAUST-MAKKAH779/2020\|EPI_ISL_ | ···················· | ···················· | ···················· | ···················· | ···················· | ···················· | | |
| F | 4782 hCoV-19/SaudiArabia/KAUST-MAKKAH781/2020\|EPI_ISL_5 | ···················· | ·A·················· | ···················· | ···················· | ···················· | ··A················· | | |
| F | 4783 hCoV-19/SaudiArabia/KAUST-MAKKAH782/2020\|EPI_ISL_5 | ···················· | ···················· | ···················· | ···················· | ···················· | ···················· | | |
| F | 4784 hCoV-19/Saudi Arabia/KAUST-MAKKAH783/2020\|EPI_ISL_ | ···················· | ·A·················· | ···················· | ···················· | ···················· | ··A················· | | |
| F | 4785 hCoV-19/SaudiArabia/KAUST-MAKKAH784/2020\|EPI_ISL_5 | ···················· | ···················· | ···················· | ···················· | ···················· | ···················· | | |
| F | 4786 hCoV-19/SaudiArabia/KAUST-MAKKAH785/2020\|EPI_ISL_5 | ···················· | ·A·················· | ···················· | ···················· | ···················· | ··A················· | | |
| F | 4787 hCoV-19/Saudi Arabia/KAUST-MAKKAH786/2020\|EPI_ISL_ | ···················· | ···················· | ···················· | ···················· | ···················· | ···················· | | |
| F | 4788 hCoV-19/SaudiArabia/KAUST-MAKKAH802/2020\|EPI_ISL_5 | ···················· | ···················· | ···················· | ···················· | ···················· | ···················· | | |
| F | 4789 hCoV-19/SaudiArabia/KAUST-MAKKAH808/2020\|EPI_ISL_5 | ···················· | ···················· | ···················· | ···················· | ···················· | ···················· | | |
| F | 4790 hCoV-19/SaudiArabia/KAUST-MAKKAH809/2020\|EPI_ISL_5 | ···················· | ···················· | ···················· | ···················· | ···················· | ···················· | | |
| F | 4791 hCoV-19/SaudiArabia/KAUST-MAKKAH810/2020\|EPI_ISL_5 | ···················· | ···················· | ···················· | ···················· | ···················· | ···················· | | |
| F | 4792 hCoV-19/SaudiArabia/KAUST-MAKKAH812/2020\|EPI_ISL_5 | ···················· | ···················· | ···················· | ···················· | ···················· | ···················· | | |
| F | 4793 hCoV-19/SaudiArabia/KAUST-MAKKAH813/2020\|EPI_ISL_5 | ···················· | ···················· | ···················· | ···················· | ···················· | ···················· | | |
| F | 4794 hCoV-19/SaudiArabia/KAUST-MAKKAH814/2020\|EPI_ISL_5 | ···················· | ···················· | ···················· | ···················· | ···················· | ···················· | | |
| F | 4795 hCoV-19/SaudiArabia/KAUST-MAKKAH815/2020\|EPI_ISL_5 | ···················· | ···················· | ···················· | ···················· | ···················· | ···················· | | |
| F | 4796 hCoV-19/USA/FL-9F97/2020\|EPI_ISL_513265\|2020-04-24 | ···················· | ···················· | ···················· | ···················· | ···················· | ···················· | | |
| F | 4797 hCoV-19/USA/FL-A9GT/2020\|EPI_ISL_513266\|2020-06-20 | ···················· | ···················· | ···················· | ···················· | ···················· | ···················· | | |
| F | 4798 hCoV-19/USA/FL-AB4M/2020\|EPI_ISL_513267\|2020-06-23 | ···················· | ···················· | ···················· | ········C·---·T·AA· | ···················· | ···················· | 1 | 1 |
| F | 4799 hCoV-19/USA/FL-ABAN/2020\|EPI_ISL_513268\|2020-06-23 | ···················· | ···················· | ···················· | ···················· | ···················· | ···················· | | |
| F | 4800 hCoV-19/USA/FL-ACYG/2020\|EPI_ISL_513269\|2020-06-26 | ···················· | ·············A······ | ···················· | ···················· | ···················· | ···················· | | |
| F | 4801 hCoV-19/USA/FL-ACYH/2020\|EPI_ISL_513270\|2020-06-26 | ···················· | ·············A······ | ···················· | ···················· | ···················· | ···················· | | |
| F | 4802 hCoV-19/USA/FL-AD68/2020\|EPI_ISL_513271\|2020-06-26 | ···················· | ···················· | ···················· | ···················· | ···················· | ··················C· | 1 | |
| F | 4803 hCoV-19/USA/FL-ADA1/2020\|EPI_ISL_513272\|2020-06-26 | ···················· | ···················· | ···················· | ···················· | ···················· | ···················· | | |
| F | 4804 hCoV-19/USA/FL-ADA3/2020\|EPI_ISL_513273\|2020-06-26 | ···················· | ···················· | ···················· | ···················· | ···················· | ···················· | | |
| F | 4805 hCoV-19/USA/FL-ADNH/2020\|EPI_ISL_513274\|2020-06-25 | ···················· | ···················· | ···················· | ···················· | ···················· | ···················· | | |
| F | 4806 hCoV-19/USA/FL-ADNP/2020\|EPI_ISL_513275\|2020-06-26 | ···················· | ···················· | ···················· | ···················· | ···················· | ···················· | | |
| F | 4807 hCoV-19/USA/FL-ADNQ/2020\|EPI_ISL_513276\|2020-06-27 | ···················· | ···················· | ···················· | ···················· | ···················· | ···················· | | |
| F | 4808 hCoV-19/USA/FL-ADNR/2020\|EPI_ISL_513277\|2020-06-26 | ···················· | ···················· | ···················· | ···················· | ···················· | ···················· | | |
| F | 4809 hCoV-19/USA/FL-ADPH/2020\|EPI_ISL_513278\|2020-06-28 | ···················· | ···················· | ···················· | ···················· | ···················· | ···················· | | |
| F | 4810 hCoV-19/USA/FL-ADPJ/2020\|EPI_ISL_513279\|2020-06-28 | ···················· | ···················· | ···················· | ···················· | ···················· | ···················· | | |
| F | 4811 hCoV-19/USA/FL-AFFD/2020\|EPI_ISL_513281\|2020-06-29 | ···················· | ···················· | ···················· | ···················· | ···················· | ···················· | | |
| F | 4812 hCoV-19/USA/FL-AFFG/2020\|EPI_ISL_513282\|2020-06-29 | ···················· | ···················· | ···················· | ···················· | ···················· | ···················· | | |
| F | 4813 hCoV-19/USA/FL-AFFK/2020\|EPI_ISL_513283\|2020-06-29 | ···················· | ···················· | ···················· | ···················· | ···················· | ···················· | | |
| F | 4814 hCoV-19/USA/FL-AFQQ/2020\|EPI_ISL_513284\|2020-06-29 | ···················· | ···················· | ···················· | ···················· | ···················· | ···················· | | |
| F | 4815 hCoV-19/USA/FL-AG7J/2020\|EPI_ISL_513285\|2020-06-30 | ···················· | ···················· | ···················· | ···················· | ···················· | ···················· | | |
| F | 4816 hCoV-19/USA/FL-AH8N/2020\|EPI_ISL_513286\|2020-06-30 | ···················· | ···················· | ···················· | ···················· | ···················· | ···················· | | |
| F | 4817 hCoV-19/USA/FL-AJ50/2020\|EPI_ISL_513287\|2020-07-04 | ···················· | ···················· | ···················· | ···················· | ···················· | ···················· | | |
| F | 4818 hCoV-19/USA/FL-AJ53/2020\|EPI_ISL_513288\|2020-07-04 | ···················· | ···················· | ···················· | ···················· | ···················· | ···················· | | |
| F | 4819 hCoV-19/USA/FL-AJ5C/2020\|EPI_ISL_513289\|2020-07-05 | ···················· | ···················· | ···················· | ···················· | ···················· | ···················· | | |
| F | 4820 hCoV-19/USA/FL-AJ5H/2020\|EPI_ISL_513290\|2020-07-04 | ···················· | ···················· | ···················· | ···················· | ···················· | ···················· | | |
| F | 4821 hCoV-19/USA/FL-AJ5U/2020\|EPI_ISL_513291\|2020-07-05 | ···················· | ···················· | ···················· | ···················· | ···················· | ···················· | | |
| F | 4822 hCoV-19/USA/PR-AGN5/2020\|EPI_ISL_513292\|2020-06-29 | ···················· | ···················· | ···················· | ···················· | ···················· | ···················· | | |
| F | 4823 hCoV-19/USA/PR-AHAA/2020\|EPI_ISL_513293\|2020-06-30 | ···················· | ···················· | ···················· | ···················· | ···················· | ···················· | | |
| F | 4825 hCoV-19/Wales/PHWC-169A60/2020\|EPI_ISL_514571\|2020 | ···················· | ···················· | ···················· | ···················· | ···················· | ···················· | | |
| F | 4826 hCoV-19/Wales/PHWC-169A7F/2020\|EPI_ISL_514572\|2020 | ···················· | ···················· | ···················· | ···················· | ···················· | ···················· | | |
| F | 4827 hCoV-19/Wales/PHWC-169A8E/2020\|EPI_ISL_514573\|2020 | ···················· | ···················· | ···················· | ···················· | NNN················· | ···················· | | 1 |
| F | 4830 hCoV-19/Wales/PHWC-169ABB/2020\|EPI_ISL_514576\|2020 | ···················· | ···················· | ···················· | ···················· | ···················· | ···················· | | |
| F | 4831 hCoV-19/Wales/PHWC-169ACA/2020\|EPI_ISL_514577\|2020 | ···················· | ···················· | ···················· | ···················· | ···················· | ···················· | | |
| F | 4832 hCoV-19/Wales/PHWC-169AD9/2020\|EPI_ISL_514578\|2020 | ···················· | ···················· | ···················· | ···················· | ···················· | ···················· | | |
| F | 4833 hCoV-19/Wales/PHWC-169AE8/2020\|EPI_ISL_514579\|2020 | ···················· | ···················· | ···················· | ···················· | ···················· | ···················· | | |
| F | 4834 hCoV-19/Bangladesh/NGRI-NSTU-16/2020\|EPI_ISL_51458 | ···················· | ···················· | ···················· | ···················· | ···················· | ···················· | | |
| F | 4835 hCoV-19/India/GJ-GBRC341/2020\|EPI_ISL_514581\|2020- | ···················· | ···················· | ···················· | ···················· | ···················· | ···················· | | |
| F | 4836 hCoV-19/India/GJ-GBRC342a/2020\|EPI_ISL_514582\|2020 | ···················· | ·A·················· | ···················· | ···················· | ···················· | ··A················· | | |
| F | 4837 hCoV-19/India/GJ-GBRC342b/2020\|EPI_ISL_514583\|2020 | ···················· | ·A·················· | ···················· | ···················· | ···················· | ··A················· | | |
| F | 4838 hCoV-19/India/GJ-GBRC343a/2020\|EPI_ISL_514584\|2020 | ···················· | ·A·················· | ···················· | ···················· | ···················· | ··A················· | | |
| F | 4839 hCoV-19/India/GJ-GBRC343b/2020\|EPI_ISL_514585\|2020 | ···················· | ·A·················· | ···················· | ···················· | ···················· | ··A················· | | |
| F | 4840 hCoV-19/India/GJ-GBRC344/2020\|EPI_ISL_514586\|2020- | ···················· | ·A·················· | ···················· | ···················· | ···················· | ··A················· | | |
| F | 4841 hCoV-19/India/GJ-GBRC345/2020\|EPI_ISL_514587\|2020- | ···················· | ·A·················· | ···················· | ···················· | ···················· | ··A················· | | |
| F | 4842 hCoV-19/India/GJ-GBRC346/2020\|EPI_ISL_514588\|2020- | ···················· | ·A·················· | ···················· | ···················· | ···················· | ··A················· | | |
| F | 4843 hCoV-19/India/GJ-GBRC347a/2020\|EPI_ISL_514589\|2020 | ···················· | ·A·················· | ···················· | ···················· | ···················· | ··A················· | | |
| F | 4844 hCoV-19/India/GJ-GBRC347b/2020\|EPI_ISL_514590\|2020 | ···················· | ·A·················· | ···················· | ···················· | ···················· | ··A················· | | |
| F | 4845 hCoV-19/India/GJ-GBRC348a/2020\|EPI_ISL_514591\|2020 | ···················· | ·A·················· | ···················· | ···················· | ···················· | ··A················· | | |
| F | 4846 hCoV-19/India/GJ-GBRC348b/2020\|EPI_ISL_514592\|2020 | ···················· | ·A·················· | ···················· | ···················· | ···················· | ··A················· | | |
| F | 4847 hCoV-19/India/GJ-GBRC349/2020\|EPI_ISL_514593\|2020- | ···················· | ··········A········· | ···················· | ···················· | ···················· | ···················· | | |
| F | 4848 hCoV-19/India/GJ-GBRC350a/2020\|EPI_ISL_514594\|2020 | ···················· | ·A·················· | ···················· | ···················· | ···················· | ··A················· | | |
| F | 4849 hCoV-19/India/GJ-GBRC350b/2020\|EPI_ISL_514595\|2020 | ···················· | ·A·················· | ···················· | ···················· | ···················· | ··A················· | | |
| F | 4850 hCoV-19/India/GJ-GBRC351a/2020\|EPI_ISL_514596\|2020 | ···················· | ·A·················· | ···················· | ···················· | ···················· | ··A················· | | |
| F | 4851 hCoV-19/India/GJ-GBRC351b/2020\|EPI_ISL_514597\|2020 | ···················· | ·A·················· | ···················· | ···················· | ···················· | ··A················· | | |
| F | 4852 hCoV-19/India/GJ-GBRC352a/2020\|EPI_ISL_514598\|2020 | ···················· | ·A·················· | ···················· | ···················· | ···················· | ···················· | | |
| F | 4853 hCoV-19/India/GJ-GBRC352b/2020\|EPI_ISL_514599\|2020 | ···················· | ···················· | ···················· | ···················· | ···················· | ···················· | | |
| F | 4854 hCoV-19/South Africa/KRISP-K001329/2020\|EPI_ISL_51 | ···················· | ···················· | ···················· | ···················· | ···················· | ···················· | | |

| | | | | | | | | |
|---|---|---|---|---|---|---|---|---|
| F | 4855 | hCoV-19/South Africa/KRISP-K001330/2020\|EPI_ISL_51 | ···················· | ···················· | ····················· | ···················· | ···················· | ···················· |
| F | 4856 | hCoV-19/South Africa/KRISP-K001331/2020\|EPI_ISL_51 | ···················· | ···················· | ····················· | ···················· | ···················· | ···················· |
| F | 4857 | hCoV-19/South Africa/KRISP-K001332/2020\|EPI_ISL_51 | ···················· | ······A············· | ····················· | ···················· | ···················· | ···················· |
| F | 4858 | hCoV-19/South Africa/KRISP-K001333/2020\|EPI_ISL_51 | ···················· | ···················· | ····················· | ···················· | ···················· | ···················· |
| F | 4859 | hCoV-19/South Africa/KRISP-K001334/2020\|EPI_ISL_51 | ···················· | ···················· | ····················· | ···················· | ···················· | ···················· |
| F | 4860 | hCoV-19/South Africa/KRISP-K001335/2020\|EPI_ISL_51 | ···················· | ···················· | ····················· | ···················· | ···················· | ···················· |
| F | 4861 | hCoV-19/South Africa/KRISP-K001336/2020\|EPI_ISL_51 | ···················· | ···················· | ····················· | ···················· | ···················· | ···················· |
| F | 4862 | hCoV-19/South Africa/KRISP-K001337/2020\|EPI_ISL_51 | ···················· | ···················· | ····················· | ···················· | ···················· | ···················· |
| F | 4863 | hCoV-19/South Africa/KRISP-K001338/2020\|EPI_ISL_51 | ···················· | ···················· | ····················· | ···················· | ···················· | ···················· |
| F | 4864 | hCoV-19/South Africa/KRISP-K001339/2020\|EPI_ISL_51 | ···················· | ···················· | ····················· | ···················· | ···················· | ···················· |
| F | 4865 | hCoV-19/USA/WA-S1777/2020\|EPI_ISL_496203\|2020-06-0 | ···················· | ···················· | ····················· | ···················· | ···················· | ···················· |
| F | 4866 | hCoV-19/USA/WA-S1808/2020\|EPI_ISL_496234\|2020-06-0 | ···················· | ···················· | ····················· | ···················· | ···················· | ···················· |
| F | 4867 | hCoV-19/USA/WA-S1809/2020\|EPI_ISL_496235\|2020-06-0 | ···················· | ···················· | ····················· | ···················· | ···················· | ···················· |
| F | 4868 | hCoV-19/USA/WA-S1810/2020\|EPI_ISL_496236\|2020-06-0 | ···················· | ···················· | ····················· | ···················· | ···················· | ···················· |
| F | 4869 | hCoV-19/USA/WA-S1811/2020\|EPI_ISL_496237\|2020-06-0 | ···················· | ···················· | ····················· | ···················· | ···················· | ···················· |
| F | 4870 | hCoV-19/USA/WA-S1812/2020\|EPI_ISL_496238\|2020-05-2 | ···················· | ···················· | ····················· | ···················· | ···················· | ···················· |
| F | 4871 | hCoV-19/USA/WA-S1813/2020\|EPI_ISL_496239\|2020-06-0 | ···················· | ···················· | ····················· | ···················· | ···················· | ···················· |
| F | 4872 | hCoV-19/USA/WA-S1814/2020\|EPI_ISL_496240\|2020-06-0 | ···················· | ···················· | ····················· | ···················· | ···················· | ···················· |
| F | 4873 | hCoV-19/USA/WA-S1815/2020\|EPI_ISL_496241\|2020-06-0 | ···················· | ···················· | ····················· | ···················· | ···················· | ···················· |
| F | 4874 | hCoV-19/USA/WA-S1816/2020\|EPI_ISL_496242\|2020-06-0 | ···················· | ···················· | ····················· | ···················· | ···················· | ···················· |
| F | 4875 | hCoV-19/USA/WA-S1817/2020\|EPI_ISL_496243\|2020-06-0 | ···················· | ···················· | ····················· | ···················· | ···················· | ···················· |
| F | 4876 | hCoV-19/USA/WA-S1818/2020\|EPI_ISL_496244\|2020-06-0 | ···················· | ···················· | ····················· | ···················· | ···················· | ···················· |
| F | 4877 | hCoV-19/USA/WA-S1819/2020\|EPI_ISL_496245\|2020-06-0 | ···················· | ···················· | ····················· | ···················· | ···················· | ···················· |
| F | 4878 | hCoV-19/USA/WA-S1820/2020\|EPI_ISL_496246\|2020-06-0 | ···················· | ···················· | ····················· | ···················· | ···················· | ···················· |
| F | 4879 | hCoV-19/USA/WA-S1822/2020\|EPI_ISL_496248\|2020-06-0 | ·T·················· | ···················· | ····················· | ···················· | ···················· | ···················· |
| F | 4880 | hCoV-19/USA/WA-S1823/2020\|EPI_ISL_496249\|2020-06-0 | ·T·················· | ···················· | ····················· | ···················· | ···················· | ···················· |
| F | 4881 | hCoV-19/USA/WA-S1824/2020\|EPI_ISL_496250\|2020-06-0 | ···················· | ···················· | ····················· | ···················· | ···················· | ···················· |
| F | 4882 | hCoV-19/USA/WA-S1826/2020\|EPI_ISL_496252\|2020-06-0 | ···················· | ···················· | ····················· | ···················· | ···················· | ···················· |
| F | 4883 | hCoV-19/USA/WA-S1827/2020\|EPI_ISL_496253\|2020-06-0 | ···················· | ···················· | ····················· | ···················· | ···················· | ···················· |
| F | 4884 | hCoV-19/USA/WA-S1829/2020\|EPI_ISL_496255\|2020-05-2 | ···················· | ···················· | ····················· | ···················· | ···················· | ···················· |
| F | 4885 | hCoV-19/USA/WA-S1830/2020\|EPI_ISL_496256\|2020-05-2 | ···················· | ···················· | ····················· | ···················· | ···················· | ···················· |
| F | 4886 | hCoV-19/USA/WA-S1831/2020\|EPI_ISL_496257\|2020-05-2 | ···················· | ···················· | ····················· | ···················· | ···················· | ···················· |
| F | 4887 | hCoV-19/USA/WA-S1832/2020\|EPI_ISL_496258\|2020-05-2 | ···················· | ···················· | ····················· | ···················· | ···················· | ···················· |
| F | 4888 | hCoV-19/USA/WA-S1833/2020\|EPI_ISL_496259\|2020-05-2 | ···················· | C··················· | ····················· | ···················· | ···················· | ···················· |
| F | 4889 | hCoV-19/USA/WA-S1834/2020\|EPI_ISL_496260\|2020-05-2 | ···················· | C··················· | ····················· | ···················· | ···················· | ···················· |
| F | 4890 | hCoV-19/USA/WA-S1835/2020\|EPI_ISL_496261\|2020-05-2 | ···················· | ···················· | ····················· | ···················· | ···················· | ···················· |
| F | 4891 | hCoV-19/USA/WA-S1837/2020\|EPI_ISL_496263\|2020-05-2 | ···················· | ···················· | ····················· | ···················· | ···················· | ···················· |
| F | 4892 | hCoV-19/USA/WA-S1838/2020\|EPI_ISL_496264\|2020-05-2 | ···················· | ···················· | ····················· | ···················· | ···················· | ···················· |
| F | 4893 | hCoV-19/USA/WA-S1839/2020\|EPI_ISL_496265\|2020-05-2 | ···················· | ···················· | ····················· | ···················· | ···················· | ···················· |
| F | 4894 | hCoV-19/USA/WA-S1840/2020\|EPI_ISL_496266\|2020-05-2 | ···················· | ···················· | ····················· | ···················· | ···················· | ···················· |
| F | 4895 | hCoV-19/USA/WA-S1841/2020\|EPI_ISL_496267\|2020-05-2 | ···················· | ···················· | ····················· | ···················· | ···················· | ···················· |
| F | 4896 | hCoV-19/USA/WA-S1842/2020\|EPI_ISL_496268\|2020-05-2 | ···················· | ···················· | ····················· | ···················· | ···················· | ···················· |
| F | 4897 | hCoV-19/USA/WA-S1843/2020\|EPI_ISL_496269\|2020-05-2 | ···················· | ···················· | ····················· | ···················· | ···················· | ···················· |
| F | 4898 | hCoV-19/USA/WA-S1844/2020\|EPI_ISL_496270\|2020-05-2 | ···················· | ···················· | ····················· | ···················· | ···················· | ···················· |
| F | 4899 | hCoV-19/USA/WA-S1845/2020\|EPI_ISL_496271\|2020-05-2 | ···················· | ···················· | ····················· | ···················· | ···················· | ···················· |
| F | 4900 | hCoV-19/USA/WA-S1846/2020\|EPI_ISL_496272\|2020-05-2 | ···················· | ···················· | ····················· | ···················· | ···················· | ···················· |
| F | 4901 | hCoV-19/USA/WA-S1847/2020\|EPI_ISL_496273\|2020-05-2 | ···················· | ···················· | ····················· | ···················· | ···················· | ···················· |
| F | 4902 | hCoV-19/USA/WA-S1848/2020\|EPI_ISL_496274\|2020-05-2 | ···················· | ···················· | ····················· | ···················· | ···················· | ···················· |
| F | 4903 | hCoV-19/USA/WA-S1849/2020\|EPI_ISL_496275\|2020-05-2 | ···················· | ···················· | ····················· | ···················· | ···················· | ···················· |
| F | 4904 | hCoV-19/USA/WA-S1850/2020\|EPI_ISL_496276\|2020-05-2 | ···················· | ···················· | ····················· | ···················· | ···················· | ···················· |
| F | 4905 | hCoV-19/USA/WA-S1851/2020\|EPI_ISL_496277\|2020-05-2 | ···················· | ···················· | ····················· | ···················· | ···················· | ···················· |
| F | 4906 | hCoV-19/USA/WA-S1852/2020\|EPI_ISL_496278\|2020-05-2 | ···················· | ···················· | ····················· | ···················· | ···················· | ···················· |
| F | 4907 | hCoV-19/USA/WA-S1854/2020\|EPI_ISL_496280\|2020-05-2 | ···················· | ···················· | ····················· | ···················· | ···················· | ···················· |
| F | 4908 | hCoV-19/USA/WA-S1855/2020\|EPI_ISL_496281\|2020-05-2 | ···················· | ···················· | ····················· | ···················· | ···················· | ···················· |
| F | 4909 | hCoV-19/USA/WA-S1856/2020\|EPI_ISL_496282\|2020-05-2 | ···················· | ···················· | ····················· | ···················· | ···················· | ···················· |
| F | 4910 | hCoV-19/USA/WA-S1857/2020\|EPI_ISL_496283\|2020-05-2 | ···················· | ···················· | ····················· | ···················· | ···················· | ···················· |
| F | 4911 | hCoV-19/USA/WA-S1859/2020\|EPI_ISL_496285\|2020-05-2 | ···················· | ···················· | ····················· | ···················· | ···················· | ···················· |
| F | 4912 | hCoV-19/USA/WA-S1860/2020\|EPI_ISL_496286\|2020-05-2 | ···················· | ···················· | ····················· | ···················· | ···················· | ···················· |
| F | 4913 | hCoV-19/USA/WA-S1861/2020\|EPI_ISL_496287\|2020-05-2 | ···················· | ···················· | ····················· | ···················· | ···················· | ···················· |
| F | 4914 | hCoV-19/USA/WA-S1862/2020\|EPI_ISL_496288\|2020-05-2 | ···················· | ···················· | ····················· | ···················· | ···················· | ···················· |
| F | 4915 | hCoV-19/USA/WA-S1863/2020\|EPI_ISL_496289\|2020-05-2 | ···················· | ···················· | ····················· | ···················· | ···················· | ···················· |
| F | 4916 | hCoV-19/USA/WA-S1864/2020\|EPI_ISL_496290\|2020-05-2 | ···················· | ···················· | ····················· | ···················· | ···················· | ···················· |
| F | 4917 | hCoV-19/USA/WA-S1865/2020\|EPI_ISL_496291\|2020-05-2 | ···················· | ···················· | ····················· | ···················· | ···················· | ···················· |
| F | 4918 | hCoV-19/USA/WA-S1866/2020\|EPI_ISL_496292\|2020-05-2 | ···················· | ···················· | ····················· | ···················· | ···················· | ···················· |
| F | 4919 | hCoV-19/USA/WA-S1868/2020\|EPI_ISL_496294\|2020-05-2 | ···················· | ···················· | ····················· | ···················· | ···················· | ···················· |
| F | 4920 | hCoV-19/USA/WA-S1869/2020\|EPI_ISL_496295\|2020-05-2 | ···················· | ···················· | ····················· | ···················· | ···················· | ···················· |
| F | 4921 | hCoV-19/USA/WA-S1870/2020\|EPI_ISL_496296\|2020-05-2 | ···················· | ···················· | ····················· | ···················· | ···················· | ···················· |
| F | 4922 | hCoV-19/USA/WA-S1871/2020\|EPI_ISL_496297\|2020-05-2 | ···················· | ···················· | ····················· | ···················· | ···················· | ···················· |
| F | 4923 | hCoV-19/USA/WA-S1872/2020\|EPI_ISL_496298\|2020-05-2 | ···················· | ···················· | ····················· | ···················· | ···················· | ···················· |
| F | 4924 | hCoV-19/USA/WA-S1873/2020\|EPI_ISL_496299\|2020-05-2 | ···················· | ···················· | ····················· | ···················· | ···················· | ···················· |
| F | 4925 | hCoV-19/USA/WA-S2107/2020\|EPI_ISL_497535\|2020-06-0 | ···················· | ···················· | ····················· | ···················· | ···················· | ···················· |
| F | 4926 | hCoV-19/USA/WA-S2137/2020\|EPI_ISL_497565\|2020-06-0 | ···················· | ···················· | ····················· | ···················· | ···················· | ···················· |
| F | 4927 | hCoV-19/USA/WA-S2139/2020\|EPI_ISL_497567\|2020-06-0 | ···················· | ···················· | ····················· | ···················· | ···················· | ···················· |
| F | 4928 | hCoV-19/USA/WA-S2141/2020\|EPI_ISL_497569\|2020-06-0 | ···················· | ···················· | ····················· | ···················· | ···················· | ···················· |
| F | 4929 | hCoV-19/USA/WA-S2142/2020\|EPI_ISL_497570\|2020-06-0 | ···················· | ·A·················· | ····················· | ···················· | ···················· | ···················· |
| F | 4930 | hCoV-19/USA/WA-S2143/2020\|EPI_ISL_497571\|2020-06-0 | ···················· | ···················· | ····················· | ···················· | ···················· | ···················· |
| F | 4931 | hCoV-19/USA/WA-S2145/2020\|EPI_ISL_497573\|2020-06-0 | ···················· | ···················· | ····················· | ···················· | ···················· | ···················· |
| F | 4932 | hCoV-19/USA/WA-S2146/2020\|EPI_ISL_497574\|2020-06-0 | ···················· | ···················· | ····················· | ···················· | ···················· | ···················· |
| F | 4933 | hCoV-19/USA/WA-S2147/2020\|EPI_ISL_497575\|2020-06-0 | ···················· | ···················· | ····················· | ···················· | ···················· | ···················· |
| F | 4934 | hCoV-19/USA/WA-S2148/2020\|EPI_ISL_497576\|2020-06-0 | ···················· | ···················· | ····················· | ···················· | ···················· | ···················· |

| | | | | | | | | |
|---|---|---|---|---|---|---|---|---|
| F | 4935 | hCoV-19/USA/WA-S2149/2020\|EPI_ISL_497577\|2020-06-0 | ···················· | ···················· | ···················· | ···················· | ···················· | ···················· |
| F | 4936 | hCoV-19/USA/WA-S2150/2020\|EPI_ISL_497578\|2020-06-0 | ···················· | ···················· | ···················· | ···················· | ···················· | ···················· |
| F | 4937 | hCoV-19/USA/WA-S2151/2020\|EPI_ISL_497579\|2020-06-0 | ···················· | ···················· | ···················· | ···················· | ···················· | ···················· |
| F | 4938 | hCoV-19/USA/WA-S2152/2020\|EPI_ISL_497580\|2020-06-0 | ···················· | ···················· | ···················· | ···················· | ···················· | ···················· |
| F | 4939 | hCoV-19/USA/WA-S2153/2020\|EPI_ISL_497581\|2020-06-0 | ···················· | ···················· | ···················· | ···················· | ···················· | ···················· |
| F | 4940 | hCoV-19/USA/WA-S2154/2020\|EPI_ISL_497582\|2020-06-0 | ···················· | ···················· | ···················· | ···················· | ···················· | ···················· |
| F | 4941 | hCoV-19/USA/WA-S2155/2020\|EPI_ISL_497583\|2020-06-0 | ···················· | ···················· | ···················· | ···················· | ···················· | ···················· |
| F | 4942 | hCoV-19/USA/WA-S2156/2020\|EPI_ISL_497584\|2020-06-0 | ···················· | ···················· | ···················· | ···················· | ···················· | ···················· |
| F | 4943 | hCoV-19/USA/WA-S2157/2020\|EPI_ISL_497585\|2020-06-0 | ···················· | ···················· | ···················· | ···················· | ···················· | ···················· |
| F | 4944 | hCoV-19/USA/WA-S2158/2020\|EPI_ISL_497586\|2020-06-0 | ···················· | ···················· | ···················· | ···················· | ···················· | ···················· |
| F | 4945 | hCoV-19/USA/WA-S2159/2020\|EPI_ISL_497587\|2020-06-0 | ···················· | ···················· | ···················· | ···················· | ···················· | ···················· |
| F | 4946 | hCoV-19/USA/WA-S2160/2020\|EPI_ISL_497588\|2020-06-0 | ···················· | ···················· | ···················· | ···················· | ···················· | ···················· |
| F | 4947 | hCoV-19/USA/WA-S2161/2020\|EPI_ISL_497589\|2020-06-0 | ···················· | ···················· | ···················· | ···················· | ···················· | ···················· |
| F | 4948 | hCoV-19/USA/WA-S2162/2020\|EPI_ISL_497590\|2020-06-0 | ···················· | ···················· | ···················· | ···················· | ···················· | ···················· |
| F | 4949 | hCoV-19/USA/WA-S2163/2020\|EPI_ISL_497591\|2020-06-0 | ···················· | ···················· | ···················· | ···················· | ···················· | ···················· |
| F | 4950 | hCoV-19/USA/WA-S2164/2020\|EPI_ISL_497592\|2020-06-0 | ···················· | ···················· | ···················· | ···················· | ···················· | ···················· |
| F | 4951 | hCoV-19/USA/WA-S2165/2020\|EPI_ISL_497593\|2020-06-0 | ···················· | ···················· | ···················· | ···················· | ···················· | ···················· |
| F | 4952 | hCoV-19/USA/WA-S2166/2020\|EPI_ISL_497594\|2020-06-0 | ···················· | ···················· | ···················· | ···················· | ···················· | ···················· |
| F | 4953 | hCoV-19/USA/WA-S2167/2020\|EPI_ISL_497595\|2020-06-0 | ···················· | ···················· | ···················· | ···················· | ···················· | ···················· |
| F | 4954 | hCoV-19/USA/WA-S2168/2020\|EPI_ISL_497596\|2020-06-0 | ···················· | ···················· | ···················· | ···················· | ···················· | ···················· |
| F | 4955 | hCoV-19/USA/WA-S2170/2020\|EPI_ISL_497598\|2020-06-0 | ···················· | ···················· | ···················· | ···················· | ···················· | ···················· |
| F | 4956 | hCoV-19/USA/WA-S2171/2020\|EPI_ISL_497599\|2020-06-0 | ···················· | ···················· | ···················· | ···················· | ···················· | ···················· |
| F | 4957 | hCoV-19/USA/WA-UW-11457/2020\|EPI_ISL_501100\|2020-0 | ···················· | ···················· | ···················· | ···················· | ···················· | ···················· |
| F | 4958 | hCoV-19/USA/WA-UW-11538/2020\|EPI_ISL_501101\|2020-0 | ···················· | ···················· | ···················· | ···················· | ···················· | ···················· |
| F | 4959 | hCoV-19/USA/WA-UW-11636/2020\|EPI_ISL_501102\|2020-0 | ···················· | ···················· | ···················· | ···················· | ···················· | ···················· |
| F | 4960 | hCoV-19/USA/WA-UW-11829/2020\|EPI_ISL_501103\|2020-0 | ···················· | ···················· | ···················· | ···················· | ···················· | ···················· |
| F | 4961 | hCoV-19/USA/WA-UW-11832/2020\|EPI_ISL_501104\|2020-0 | ···················· | ·A·················· | ···················· | ···················· | ···················· | ···················· |
| F | 4962 | hCoV-19/USA/WA-UW-11918/2020\|EPI_ISL_501105\|2020-0 | ···················· | ···················· | ···················· | ···················· | ···················· | ···················· |
| F | 4963 | hCoV-19/USA/WA-UW-12027/2020\|EPI_ISL_501106\|2020-0 | ···················· | ···················· | ···················· | ···················· | ···················· | ···················· |
| F | 4964 | hCoV-19/USA/WA-UW-12149/2020\|EPI_ISL_501107\|2020-0 | ···················· | ···················· | ···················· | ···················· | ···················· | ···················· |
| F | 4965 | hCoV-19/USA/WA-UW-12264/2020\|EPI_ISL_501108\|2020-0 | ···················· | ·A·················· | ···················· | ···················· | ···················· | ···················· |
| F | 4966 | hCoV-19/USA/WA-UW-12372/2020\|EPI_ISL_501109\|2020-0 | ···················· | ···················· | ···················· | ···················· | ···················· | ···················· |
| F | 4967 | hCoV-19/USA/WA-UW-12396/2020\|EPI_ISL_501110\|2020-0 | ···················· | ···················· | ···················· | ···················· | ···················· | ···················· |
| F | 4968 | hCoV-19/USA/WA-UW-12446/2020\|EPI_ISL_501111\|2020-0 | ···················· | ·A·················· | ···················· | ···················· | ···················· | ···················· |
| F | 4969 | hCoV-19/USA/WA-UW-12596/2020\|EPI_ISL_501112\|2020-0 | ···················· | ···················· | ···················· | ···················· | ···················· | ···················· |
| F | 4970 | hCoV-19/USA/WA-UW-12612/2020\|EPI_ISL_501113\|2020-0 | ···················· | ···················· | ···················· | ···················· | ···················· | ···················· |
| F | 4971 | hCoV-19/USA/WA-UW-12689/2020\|EPI_ISL_501114\|2020-0 | ···················· | ···················· | ···················· | ···················· | ···················· | ···················· |
| F | 4972 | hCoV-19/USA/WA-UW-12705/2020\|EPI_ISL_501115\|2020-0 | ···················· | ···················· | ···················· | ···················· | ···················· | ···················· |
| F | 4973 | hCoV-19/USA/WA-UW-12706/2020\|EPI_ISL_501116\|2020-0 | ···················· | ···················· | ···················· | ···················· | ···················· | ···················· |
| F | 4974 | hCoV-19/USA/WA-UW-12727/2020\|EPI_ISL_501117\|2020-0 | ···········T········ | ···················· | ···················· | ···················· | ···················· | ···················· |
| F | 4975 | hCoV-19/USA/WA-UW-13028/2020\|EPI_ISL_501118\|2020-0 | ···················· | ···················· | ···················· | ···················· | ···················· | ···················· |
| F | 4976 | hCoV-19/USA/WA-UW-13226/2020\|EPI_ISL_501120\|2020-0 | ···················· | ···················· | ···················· | ···················· | ···················· | ···················· |
| F | 4977 | hCoV-19/USA/WA-UW-13227/2020\|EPI_ISL_501121\|2020-0 | ···················· | ···················· | ···················· | ···················· | ···················· | ···················· |
| F | 4978 | hCoV-19/USA/WA-UW-13232/2020\|EPI_ISL_501122\|2020-0 | ···················· | ···················· | ···················· | ···················· | ···················· | ···················· |
| F | 4979 | hCoV-19/USA/WA-UW-13233/2020\|EPI_ISL_501123\|2020-0 | ···················· | ···················· | ···················· | ···················· | ···················· | ···················· |
| F | 4980 | hCoV-19/USA/WA-UW-13242/2020\|EPI_ISL_501124\|2020-0 | ···················· | ···················· | ···················· | ···················· | ···················· | ···················· |
| F | 4981 | hCoV-19/USA/WA-UW-13262/2020\|EPI_ISL_501125\|2020-0 | ···················· | ·A·················· | ···················· | ···················· | ···················· | ···················· |
| F | 4982 | hCoV-19/USA/WA-UW-13264/2020\|EPI_ISL_501126\|2020-0 | ···················· | ·A·················· | ···················· | ···················· | ···················· | ···················· |
| F | 4983 | hCoV-19/USA/WA-UW-13268/2020\|EPI_ISL_501127\|2020-0 | ···················· | ·A·················· | ···················· | ···················· | ···················· | ···················· |
| F | 4984 | hCoV-19/USA/WA-UW-13274/2020\|EPI_ISL_501128\|2020-0 | ···················· | ···················· | ···················· | ···················· | ···················· | ···················· |
| F | 4985 | hCoV-19/USA/WA-UW-13288/2020\|EPI_ISL_501131\|2020-0 | ···················· | ···················· | ···················· | ···················· | ···················· | ···················· |
| F | 4986 | hCoV-19/USA/WA-UW-13291/2020\|EPI_ISL_501132\|2020-0 | ···················· | ···················· | ···················· | ···················· | ···················· | ···················· |
| F | 4987 | hCoV-19/USA/WA-UW-13293/2020\|EPI_ISL_501133\|2020-0 | ···················· | ···················· | ···················· | ···················· | ···················· | ···················· |
| F | 4988 | hCoV-19/USA/WA-UW-13300/2020\|EPI_ISL_501134\|2020-0 | ···················· | ···················· | ···················· | ···················· | ···················· | ···················· |
| F | 4989 | hCoV-19/USA/WA-UW-13306/2020\|EPI_ISL_501135\|2020-0 | ···················· | ·A·················· | ···················· | ···················· | ···················· | ···················· |
| F | 4990 | hCoV-19/USA/WA-UW-13310/2020\|EPI_ISL_501136\|2020-0 | ···················· | ···················· | ···················· | ···················· | ···················· | ···················· |
| F | 4991 | hCoV-19/USA/WA-UW-13317/2020\|EPI_ISL_501137\|2020-0 | ···················· | ·A·················· | ···················· | ···················· | ···················· | ···················· |
| F | 4992 | hCoV-19/USA/WA-UW-13321/2020\|EPI_ISL_501138\|2020-0 | ···················· | ·A·················· | ···················· | ···················· | ···················· | ···················· |
| F | 4993 | hCoV-19/Russia/StPetersburg-RII11168S/2020\|EPI_ISL | ···················· | ···················· | ···················· | ···················· | ···················· | ···················· |
| F | 4994 | hCoV-19/Russia/StPetersburg-RII11172S/2020\|EPI_ISL | ···················· | ···················· | ···················· | ···················· | ···················· | ···················· |
| F | 4995 | hCoV-19/Russia/StPetersburg-RII11183S/2020\|EPI_ISL | ···················· | ···················· | ···················· | ···················· | ···················· | ···················· |
| F | 4996 | hCoV-19/Russia/StPetersburg-RII11184S/2020\|EPI_ISL | ···················· | ···················· | ···················· | ···················· | ···················· | ···················· |
| F | 4997 | hCoV-19/Russia/StPetersburg-RII11188S/2020\|EPI_ISL | ···················· | ···················· | ···················· | ···················· | ···················· | ···················· |
| F | 4998 | hCoV-19/Russia/StPetersburg-RII11247S/2020\|EPI_ISL | ···················· | ···················· | ···················· | ···················· | ···················· | ···················· |
| F | 4999 | hCoV-19/Russia/StPetersburg-RII11250S/2020\|EPI_ISL | ···················· | ···················· | ···················· | ···················· | ···················· | ···················· |
| F | 5000 | hCoV-19/Russia/StPetersburg-RII11251S/2020\|EPI_ISL | ···················· | ···················· | ···················· | ···················· | ···················· | ···················· |
| F | 5001 | hCoV-19/Russia/StPetersburg-RII11256S/2020\|EPI_ISL | ···················· | ···················· | ···················· | ···················· | ···················· | ···················· |
| F | 5002 | hCoV-19/Russia/StPetersburg-RII11271S/2020\|EPI_ISL | ···················· | ···················· | ···················· | ···················· | ···················· | ···················· |
| F | 5003 | hCoV-19/Russia/StPetersburg-RII11273S/2020\|EPI_ISL | ···················· | ···················· | ···················· | ···················· | ···················· | ···················· |
| F | 5004 | hCoV-19/Russia/StPetersburg-RII11275S/2020\|EPI_ISL | ···················· | ···················· | ···················· | ···················· | ···················· | ···················· |
| F | 5005 | hCoV-19/Russia/StPetersburg-RII11292S/2020\|EPI_ISL | ···················· | ···················· | ···················· | ···················· | ···················· | ···················· |
| F | 5006 | hCoV-19/Russia/StPetersburg-RII11299S/2020\|EPI_ISL | ···················· | ···················· | ···················· | ···················· | ···················· | ···················· |
| F | 5007 | hCoV-19/Russia/StPetersburg-RII11306S/2020\|EPI_ISL | ···················· | ···················· | ···················· | ···················· | ···················· | ···················· |
| F | 5008 | hCoV-19/Russia/StPetersburg-RII11311S/2020\|EPI_ISL | ···················· | ···················· | ···················· | ···················· | ···················· | ···················· |
| F | 5009 | hCoV-19/Russia/StPetersburg-RII13532S/2020\|EPI_ISL | ···················· | ···················· | ···················· | ···················· | ···················· | ···················· |
| F | 5010 | hCoV-19/Russia/StPetersburg-RII14570V/2020\|EPI_ISL | ···················· | ···················· | ···················· | ···················· | ···················· | ···················· |
| F | 5011 | hCoV-19/Russia/StPetersburg-RII14691V/2020\|EPI_ISL | ···················· | ···················· | ···················· | ···················· | ···················· | ···················· |
| F | 5012 | hCoV-19/Russia/StPetersburg-RII15065S/2020\|EPI_ISL | ···················· | ···················· | ···················· | ···················· | ···················· | ···················· |
| F | 5013 | hCoV-19/Russia/StPetersburg-RII15163S/2020\|EPI_ISL | ···················· | ···················· | ···················· | ···················· | ···················· | ···················· |
| F | 5014 | hCoV-19/Russia/StPetersburg-RII15221S/2020\|EPI_ISL | ···················· | ···················· | ···················· | ···················· | ···················· | ···················· |

| | | | | | | | |
|---|---|---|---|---|---|---|---|
| F | 5015 hCoV-19/Russia/StPetersburg-RII15228S/2020\|EPI_ISL | ···················· | ···················· | ····················· | ···················· | ···················· | ···················· |
| F | 5016 hCoV-19/Russia/StPetersburg-RII15229S/2020\|EPI_ISL | ···················· | ···················· | ····················· | ···················· | ···················· | ···················· |
| F | 5017 hCoV-19/Russia/StPetersburg-RII15231S/2020\|EPI_ISL | ···················· | ···················· | ····················· | ···················· | ···················· | ···················· |
| F | 5018 hCoV-19/Russia/StPetersburg-RII15516S/2020\|EPI_ISL | ···················· | ···················· | ····················· | ···················· | ···················· | ···················· |
| F | 5019 hCoV-19/Russia/StPetersburg-RII15517S/2020\|EPI_ISL | ···················· | ···················· | ····················· | ···················· | ···················· | ···················· |
| F | 5020 hCoV-19/Russia/StPetersburg-RII15518S/2020\|EPI_ISL | ···················· | ···················· | ····················· | ···················· | ···················· | ···················· |
| F | 5021 hCoV-19/Russia/StPetersburg-RII15520S/2020\|EPI_ISL | ···················· | ···················· | ····················· | ···················· | ···················· | ···················· |
| F | 5022 hCoV-19/Russia/StPetersburg-RII15671S/2020\|EPI_ISL | ···················· | ···················· | ····················· | ···················· | ···················· | ···················· |
| F | 5023 hCoV-19/Russia/StPetersburg-RII15673S/2020\|EPI_ISL | ···················· | ···················· | ····················· | ···················· | ···················· | ···················· |
| F | 5024 hCoV-19/Russia/StPetersburg-RII15674S/2020\|EPI_ISL | ···················· | ···················· | ····················· | ···················· | ···················· | ···················· |
| F | 5025 hCoV-19/Russia/StPetersburg-RII15675S/2020\|EPI_ISL | ···················· | ···················· | ····················· | ···················· | ···················· | ···················· |
| F | 5026 hCoV-19/Russia/StPetersburg-RII15817S/2020\|EPI_ISL | ···················· | ···················· | ····················· | ···················· | ···················· | ···················· |
| F | 5027 hCoV-19/Russia/StPetersburg-RII15932V/2020\|EPI_ISL | ···················· | ···················· | ····················· | ···················· | ···················· | ···················· |
| F | 5028 hCoV-19/Russia/StPetersburg-RII15933S/2020\|EPI_ISL | ···················· | ···················· | ····················· | ···················· | ···················· | ···················· |
| F | 5029 hCoV-19/Russia/StPetersburg-RII15950V/2020\|EPI_ISL | ···················· | ···················· | ····················· | ···················· | ···················· | ···················· |
| F | 5030 hCoV-19/Russia/StPetersburg-RII16106S/2020\|EPI_ISL | ···················· | ···················· | ····················· | ···················· | ···················· | ···················· |
| F | 5031 hCoV-19/Russia/StPetersburg-RII17536S/2020\|EPI_ISL | ···················· | ·············A······ | ····················· | ···················· | ···················· | ···················· |
| F | 5032 hCoV-19/Russia/StPetersburg-RII17574S/2020\|EPI_ISL | ···················· | ···················· | ····················· | ···················· | ···················· | ···················· |
| F | 5033 hCoV-19/Russia/StPetersburg-RII17948S/2020\|EPI_ISL | ···················· | ···················· | ····················· | ···················· | ···················· | ···················· |
| F | 5034 hCoV-19/Russia/StPetersburg-RII17949S/2020\|EPI_ISL | ···················· | ···················· | ····················· | ···················· | ···················· | ···················· |
| F | 5035 hCoV-19/Russia/StPetersburg-RII21802S/2020\|EPI_ISL | ···················· | ···················· | ····················· | ···················· | ···················· | ···················· |
| F | 5036 hCoV-19/Russia/StPetersburg-RII3524VR4/2020\|EPI_IS | ···················· | ···················· | ····················· | ···················· | ···················· | ···················· |
| F | 5037 hCoV-19/Russia/StPetersburg-RII4586S/2020\|EPI_ISL_ | ···················· | ···················· | ····················· | ···················· | ···················· | ···················· |
| F | 5038 hCoV-19/Russia/StPetersburg-RII8317V/2020\|EPI_ISL_ | ···················· | ···················· | ····················· | ···················· | ···················· | ···················· |
| F | 5039 hCoV-19/Russia/StPetersburg-RII8319V/2020\|EPI_ISL_ | ···················· | ···················· | ····················· | ···················· | ···················· | ···················· |
| F | 5040 hCoV-19/Russia/StPetersburg-RII9304V/2020\|EPI_ISL_ | ···················· | ···················· | ····················· | ···················· | ···················· | ···················· |
| F | 5041 hCoV-19/Russia/SPE-RII-9364V/2020\|EPI_ISL_507296\|2 | ···················· | ···················· | ····················· | ···················· | ···················· | ···················· |
| F | 5042 hCoV-19/Russia/BA-RII-12-5800V/2020\|EPI_ISL_507297 | ···················· | ···················· | ····················· | ···················· | ···················· | ···················· |
| F | 5043 hCoV-19/Russia/BA-RII-21-5809V/2020\|EPI_ISL_507298 | ···················· | ···················· | ····················· | ···················· | ···················· | ···················· |
| F | 5044 hCoV-19/Russia/BA-RII-5-5798V/2020\|EPI_ISL_507299\| | ···················· | ···················· | ····················· | ···················· | ···················· | ···················· |
| F | 5045 hCoV-19/USA/NM-NMDOH-2020242038/2020\|EPI_ISL_51088 | ···················· | ·A·················· | ····················· | ···················· | ···················· | ···················· |
| F | 5046 hCoV-19/USA/NM-NMDOH-2020242040/2020\|EPI_ISL_51088 | ···················· | ·A·················· | ····················· | ···················· | ···················· | ···················· |
| F | 5047 hCoV-19/USA/NM-NMDOH-2020242041/2020\|EPI_ISL_51088 | ···················· | ·A·················· | ····················· | ···················· | ···················· | ···················· |
| F | 5048 hCoV-19/Portugal/PT1521/2020\|EPI_ISL_510889\|2020-0 | ···················· | ···················· | ····················· | ···················· | ···················· | ···················· |
| F | 5049 hCoV-19/Portugal/PT1522/2020\|EPI_ISL_510890\|2020-0 | ···················· | ···················· | ····················· | ···················· | ···················· | ···················· |
| F | 5050 hCoV-19/Portugal/PT0630/2020\|EPI_ISL_510891\|2020-0 | ···················· | ···················· | ····················· | ···················· | ···················· | ···················· |
| F | 5051 hCoV-19/Portugal/PT0631/2020\|EPI_ISL_510892\|2020-0 | ···················· | ···················· | ····················· | ···················· | ···················· | ···················· |
| F | 5052 hCoV-19/Portugal/PT0632/2020\|EPI_ISL_510893\|2020-0 | ···················· | ···················· | ····················· | ···················· | ···················· | ···················· |
| F | 5053 hCoV-19/Portugal/PT0635/2020\|EPI_ISL_510896\|2020-0 | ···················· | ···················· | ····················· | ···················· | ···················· | ···················· |
| F | 5054 hCoV-19/Portugal/PT0637/2020\|EPI_ISL_510898\|2020-0 | ···················· | ···················· | ····················· | ···················· | ···················· | ···················· |
| F | 5055 hCoV-19/SaudiArabia/KAUST-MADINAH1087/2020\|EPI_ISL | ···················· | ···················· | ····················· | ···················· | ···················· | ···················· |
| F | 5056 hCoV-19/SaudiArabia/KAUST-MADINAH1090/2020\|EPI_ISL | ···················· | ·A·················· | ····················· | ···················· | ···················· | ··A················· |
| F | 5057 hCoV-19/SaudiArabia/KAUST-MADINAH1093/2020\|EPI_ISL | ···················· | ·A·················· | ····················· | ···················· | ···················· | ··A················· |
| F | 5058 hCoV-19/SaudiArabia/KAUST-MADINAH1094/2020\|EPI_ISL | ···················· | ·A·················· | ····················· | ···················· | ···················· | ··A················· |
| F | 5059 hCoV-19/SaudiArabia/KAUST-MADINAH1095/2020\|EPI_ISL | ···················· | ·A·················· | ····················· | ···················· | ···················· | ··A················· |
| F | 5060 hCoV-19/SaudiArabia/KAUST-MADINAH1096/2020\|EPI_ISL | ···················· | ·A·················· | ····················· | ···················· | ···················· | ··A················· |
| F | 5061 hCoV-19/SaudiArabia/KAUST-MADINAH1099/2020\|EPI_ISL | ···················· | ·A·················· | ····················· | ···················· | ···················· | ··A················· |
| F | 5062 hCoV-19/SaudiArabia/KAUST-MADINAH1100/2020\|EPI_ISL | ···················· | ·A·················· | ····················· | ···················· | ···················· | ··A················· |
| F | 5063 hCoV-19/SaudiArabia/KAUST-MADINAH1102/2020\|EPI_ISL | ···················· | ···················· | ····················· | ···················· | ···················· | ···················· |
| F | 5064 hCoV-19/SaudiArabia/KAUST-MADINAH1103/2020\|EPI_ISL | ···················· | ···················· | ····················· | ···················· | ···················· | ···················· |
| F | 5065 hCoV-19/SaudiArabia/KAUST-MADINAH1107/2020\|EPI_ISL | ···················· | ···················· | ····················· | ···················· | ···················· | ···················· |
| F | 5066 hCoV-19/SaudiArabia/KAUST-MADINAH1108/2020\|EPI_ISL | ···················· | ···················· | ····················· | ···················· | ···················· | ···················· |
| F | 5067 hCoV-19/SaudiArabia/KAUST-MADINAH1109/2020\|EPI_ISL | ···················· | ···················· | ····················· | ···················· | ···················· | ···················· |
| F | 5068 hCoV-19/SaudiArabia/KAUST-MADINAH1110/2020\|EPI_ISL | ···················· | ···················· | ····················· | ···················· | ···················· | ···················· |
| F | 5069 hCoV-19/SaudiArabia/KAUST-MADINAH1111/2020\|EPI_ISL | ···················· | ···················· | ····················· | ···················· | ···················· | ··A················· |
| F | 5070 hCoV-19/SaudiArabia/KAUST-MADINAH1112/2020\|EPI_ISL | ···················· | ·A·················· | ····················· | ···················· | ···················· | ··A················· |
| F | 5071 hCoV-19/SaudiArabia/KAUST-MADINAH1113/2020\|EPI_ISL | ···················· | ·A·················· | ····················· | ···················· | ···················· | ··A················· |
| F | 5072 hCoV-19/SaudiArabia/KAUST-MADINAH1114/2020\|EPI_ISL | ···················· | ·A·················· | ····················· | ···················· | ···················· | ··A················· |
| F | 5073 hCoV-19/South Africa/LR00157/2020\|EPI_ISL_514424\|2 | ···················· | ···················· | ····················· | ···················· | ···················· | ···················· |
| F | 5074 hCoV-19/Serbia/KV08-15-07/2020\|EPI_ISL_514425\|2020 | ···················· | ···················· | ····················· | ···················· | ···················· | ···················· |
| F | 5075 hCoV-19/Serbia/KV52-12-05/2020\|EPI_ISL_514426\|2020 | ···················· | ···················· | ····················· | ···················· | ···················· | ···················· |
| F | 5076 hCoV-19/Serbia/KV57-12-07/2020\|EPI_ISL_514427\|2020 | ···················· | ···················· | ····················· | ···················· | ···················· | ···················· |
| F | 5077 hCoV-19/Serbia/KV65-16-07/2020\|EPI_ISL_514428\|2020 | ···················· | ···················· | ····················· | ···················· | ···················· | ···················· |
| F | 5078 hCoV-19/Serbia/NP07-15-07/2020\|EPI_ISL_514429\|2020 | ···················· | ···················· | ····················· | ···················· | ···················· | ···················· |
| F | 5079 hCoV-19/Serbia/UE06-28-04/2020\|EPI_ISL_514430\|2020 | ···················· | ···················· | ····················· | ···················· | ···················· | ···················· |
| F | 5080 hCoV-19/Serbia/UE16-22-04/2020\|EPI_ISL_514431\|2020 | ···················· | ···················· | ····················· | ···················· | ···················· | ···················· |
| F | 5081 hCoV-19/Italy/CAM-CEINGE-5/2020\|EPI_ISL_514432\|202 | ···················· | ···················· | ····················· | ···················· | ···················· | ···················· |
| F | 5082 hCoV-19/India/GJ-GBRC337a/2020\|EPI_ISL_514435\|2020 | ···················· | ···················· | ····················· | ···················· | ···················· | ···················· |
| F | 5083 hCoV-19/India/GJ-GBRC337b/2020\|EPI_ISL_514436\|2020 | ···················· | ···················· | ····················· | ···················· | ···················· | ···················· |
| F | 5084 hCoV-19/India/GJ-GBRC340/2020\|EPI_ISL_514437\|2020- | ···················· | ···················· | ····················· | ···················· | ···················· | ···················· |
| F | 5085 hCoV-19/Bangladesh/NGRI-NSTU-03/2020\|EPI_ISL_51444 | ···················· | ···················· | ····················· | ···················· | ···················· | ···················· |
| F | 5086 hCoV-19/England/CAMB-1B55CE/2020\|EPI_ISL_514443\|20 | ···················· | ···················· | ····················· | ···················· | ···················· | ···················· |
| F | 5087 hCoV-19/England/CAMB-1B592C/2020\|EPI_ISL_514445\|20 | ···················· | ···················· | ····················· | ···················· | ···················· | ···················· |
| F | 5088 hCoV-19/England/CAMB-1B5A29/2020\|EPI_ISL_514448\|20 | ···················· | ···················· | ····················· | ···················· | ···················· | ···················· |
| F | 5089 hCoV-19/England/CAMB-1B5A65/2020\|EPI_ISL_514449\|20 | ···················· | ···················· | ····················· | ···················· | ···················· | ···················· |
| F | 5090 hCoV-19/South Africa/KRISP-K002369/2020\|EPI_ISL_51 | ···················· | ···················· | ····················· | ···················· | ···················· | ···················· |
| F | 5091 hCoV-19/South Africa/KRISP-K002370/2020\|EPI_ISL_51 | ···················· | ···················· | ····················· | ···················· | ···················· | ···················· |
| F | 5092 hCoV-19/South Africa/KRISP-K002371/2020\|EPI_ISL_51 | ···················· | ···················· | ····················· | ···················· | ···················· | ···················· |
| F | 5093 hCoV-19/South Africa/KRISP-K002372/2020\|EPI_ISL_51 | ···················· | ···················· | ····················· | ···················· | ···················· | ···················· |
| F | 5094 hCoV-19/South Africa/KRISP-K002375/2020\|EPI_ISL_51 | ···················· | ···················· | ····················· | ···················· | ···················· | ···················· |

```
F 5095 hCoV-19/South Africa/KRISP-K002377/2020|EPI_ISL_51 .................... .................... .................... .................... .................... ....................
F 5096 hCoV-19/South Africa/KRISP-K002378/2020|EPI_ISL_51 .................... .................... .................... .................... .................... ....................
F 5097 hCoV-19/South Africa/KRISP-K002379/2020|EPI_ISL_51 .................... .................... .................... .................... .................... ....................
F 5098 hCoV-19/South Africa/KRISP-K002381/2020|EPI_ISL_51 .................... .................... .................... .................... .................... ....................
F 5099 hCoV-19/South Africa/KRISP-K002382/2020|EPI_ISL_51 .................... .................... .................... .................... .................... ....................
F 5100 hCoV-19/South Africa/KRISP-K002383/2020|EPI_ISL_51 .................... .................... .................... .................... .................... ....................
F 5101 hCoV-19/South Africa/KRISP-K002385/2020|EPI_ISL_51 .................... .................... .................... .................... .................... ....................
F 5102 hCoV-19/South Africa/KRISP-K002386/2020|EPI_ISL_51 .................... .................... .................... .................... .................... ....................
F 5103 hCoV-19/South Africa/KRISP-K002387/2020|EPI_ISL_51 .................... .................... .................... .................... .................... ....................
F 5104 hCoV-19/South Africa/KRISP-K002388/2020|EPI_ISL_51 .................... .................... .................... .................... .................... ....................
F 5105 hCoV-19/South Africa/KRISP-K002391/2020|EPI_ISL_51 .................... .................... .................... .................... .................... ....................
F 5106 hCoV-19/South Africa/KRISP-K002392/2020|EPI_ISL_51 .................... .................... .................... .................... .................... ....................
F 5107 hCoV-19/South Africa/KRISP-K002393/2020|EPI_ISL_51 .................... .................... .................... .................... .................... ....................
F 5108 hCoV-19/South Africa/KRISP-K002394/2020|EPI_ISL_51 .................... .................... .................... .................... .................... ....................
F 5109 hCoV-19/South Africa/KRISP-K002395/2020|EPI_ISL_51 .................... .................... .................... .................... .................... ....................
F 5110 hCoV-19/South Africa/KRISP-K002398/2020|EPI_ISL_51 .................... .................... .................... .................... .................... ....................
F 5111 hCoV-19/South Africa/KRISP-K002399/2020|EPI_ISL_51 .................... .................... .................... .................... .................... ....................
F 5112 hCoV-19/South Africa/KRISP-K002400/2020|EPI_ISL_51 .................... .................... .................... .................... .................... ....................
F 5113 hCoV-19/South Africa/KRISP-K002403/2020|EPI_ISL_51 .................... .................... .................... .................... .................... ....................
F 5114 hCoV-19/South Africa/KRISP-K002405/2020|EPI_ISL_51 .................... .................... .................... .................... .................... ....................
F 5115 hCoV-19/South Africa/KRISP-K002407/2020|EPI_ISL_51 .................... .................... .................... .................... .................... ....................
F 5116 hCoV-19/South Africa/KRISP-K002411/2020|EPI_ISL_51 .................... .................... .................... .................... .................... ....................
F 5117 hCoV-19/South Africa/KRISP-K002413/2020|EPI_ISL_51 .................... .................... .................... .................... .................... ....................
F 5118 hCoV-19/South Africa/KRISP-K002414/2020|EPI_ISL_51 .................... .................... .................... .................... .................... ....................
F 5119 hCoV-19/South Africa/KRISP-K002415/2020|EPI_ISL_51 .................... .................... .................... .................... .................... ....................
F 5120 hCoV-19/South Africa/KRISP-K002416/2020|EPI_ISL_51 .................... .................... .................... .................... .................... ....................
F 5121 hCoV-19/USA/WA-S1774/2020|EPI_ISL_496200|2020-06-0 .................... .................... .................... .................... .................... ....................
F 5122 hCoV-19/USA/WA-S1775/2020|EPI_ISL_496201|2020-06-0 .................... .................... .................... .................... .................... ....................
F 5123 hCoV-19/USA/WA-S1776/2020|EPI_ISL_496202|2020-06-0 .................... .................... .................... .................... .................... ....................
F 5124 hCoV-19/USA/WA-S1778/2020|EPI_ISL_496204|2020-06-0 .................... .................... .................... .................... .................... ....................
F 5125 hCoV-19/USA/WA-S1779/2020|EPI_ISL_496205|2020-06-0 .................... .................... .................... .................... .................... ....................
F 5126 hCoV-19/USA/WA-S1780/2020|EPI_ISL_496206|2020-06-0 .................... .................... .................... .................... .................... ....................
F 5127 hCoV-19/USA/WA-S1781/2020|EPI_ISL_496207|2020-06-0 .................... .................... .................... .................... .................... ....................
F 5128 hCoV-19/USA/WA-S1782/2020|EPI_ISL_496208|2020-06-0 .................... .................... .................... .................... .................... ....................
F 5129 hCoV-19/USA/WA-S1783/2020|EPI_ISL_496209|2020-06-0 .................... .................... .................... .................... .................... ....................
F 5130 hCoV-19/USA/WA-S1784/2020|EPI_ISL_496210|2020-06-0 .................... .................... .................... .................... .................... ....................
F 5131 hCoV-19/USA/WA-S1785/2020|EPI_ISL_496211|2020-06-0 .................... .................... .................... .................... .................... ....................
F 5132 hCoV-19/USA/WA-S1786/2020|EPI_ISL_496212|2020-06-0 .................... .................... .................... .................... .................... ....................
F 5133 hCoV-19/USA/WA-S1787/2020|EPI_ISL_496213|2020-06-0 .................... .................... .................... .................... .................... ....................
F 5134 hCoV-19/USA/WA-S1788/2020|EPI_ISL_496214|2020-06-0 .................... .................... .................... .................... .................... ....................
F 5135 hCoV-19/USA/WA-S1789/2020|EPI_ISL_496215|2020-06-0 .................... .................... .................... .................... .................... ....................
F 5136 hCoV-19/USA/WA-S1790/2020|EPI_ISL_496216|2020-06-0 .................... .................... .................... .................... .................... ....................
F 5137 hCoV-19/USA/WA-S1791/2020|EPI_ISL_496217|2020-06-0 .................... .................... .................... .................... .................... ....................
F 5138 hCoV-19/USA/WA-S1792/2020|EPI_ISL_496218|2020-06-0 .................... .................... .................... .................... .................... ....................
F 5139 hCoV-19/USA/WA-S1793/2020|EPI_ISL_496219|2020-06-0 .................... .................... .................... .................... .................... ....................
F 5140 hCoV-19/USA/WA-S1794/2020|EPI_ISL_496220|2020-06-0 .................... .................... .................... .................... .................... ....................
F 5141 hCoV-19/USA/WA-S1795/2020|EPI_ISL_496221|2020-06-0 .................... .................... .................... .................... .................... ....................
F 5142 hCoV-19/USA/WA-S1796/2020|EPI_ISL_496222|2020-06-0 .................... .................... .................... .................... .................... ....................
F 5143 hCoV-19/USA/WA-S1797/2020|EPI_ISL_496223|2020-06-0 .................... .................... .................... .................... .................... ....................
F 5144 hCoV-19/USA/WA-S1798/2020|EPI_ISL_496224|2020-06-0 .................... .................... .................... .................... .................... ....................
F 5145 hCoV-19/USA/WA-S1799/2020|EPI_ISL_496225|2020-06-0 .................... .................... .................... .................... .................... ....................
F 5146 hCoV-19/USA/WA-S1800/2020|EPI_ISL_496226|2020-06-0 .................... .................... .................... .................... .................... ....................
F 5147 hCoV-19/USA/WA-S1801/2020|EPI_ISL_496227|2020-06-0 .................... .................... .................... .................... .................... ....................
F 5148 hCoV-19/USA/WA-S1802/2020|EPI_ISL_496228|2020-06-0 .................... .................... .................... .................... .................... ....................
F 5149 hCoV-19/USA/WA-S1803/2020|EPI_ISL_496229|2020-06-0 .................... .................... .................... .................... .................... ....................
F 5150 hCoV-19/USA/WA-S1804/2020|EPI_ISL_496230|2020-06-0 .................... .................... .................... .................... .................... ....................
F 5151 hCoV-19/USA/WA-S1805/2020|EPI_ISL_496231|2020-06-0 .................... .................... .................... .................... .................... ....................
F 5152 hCoV-19/USA/WA-S1806/2020|EPI_ISL_496232|2020-06-0 .................... .................... .................... .................... .................... ....................
F 5153 hCoV-19/USA/WA-S1807/2020|EPI_ISL_496233|2020-06-0 .................... .................... .................... .................... .................... ....................
F 5154 hCoV-19/USA/WA-S2072/2020|EPI_ISL_497500|2020-06-0 .................... .................... .................... .................... .................... ....................
F 5155 hCoV-19/USA/WA-S2073/2020|EPI_ISL_497501|2020-06-0 .................... .................... .................... .................... .................... ....................
F 5156 hCoV-19/USA/WA-S2074/2020|EPI_ISL_497502|2020-06-0 .................... .................... .................... .................... .................... ....................
F 5157 hCoV-19/USA/WA-S2075/2020|EPI_ISL_497503|2020-06-0 .................... .................... .................... .................... .................... ....................
F 5158 hCoV-19/USA/WA-S2076/2020|EPI_ISL_497504|2020-06-0 .................... .................... .................... .................... .................... ....................
F 5159 hCoV-19/USA/WA-S2077/2020|EPI_ISL_497505|2020-06-0 .................... .................... .................... .................... .................... ....................
F 5160 hCoV-19/USA/WA-S2078/2020|EPI_ISL_497506|2020-06-0 .................... .................... .................... .................... .................... ....................
F 5161 hCoV-19/USA/WA-S2079/2020|EPI_ISL_497507|2020-06-0 .................... .................... .................... .................... .................... ....................
F 5162 hCoV-19/USA/WA-S2080/2020|EPI_ISL_497508|2020-06-0 .................... .................... .................... .................... .................... ....................
F 5163 hCoV-19/USA/WA-S2081/2020|EPI_ISL_497509|2020-06-0 .................... .................... .................... .................... .................... ....................
F 5164 hCoV-19/USA/WA-S2082/2020|EPI_ISL_497510|2020-06-0 .................... .................... .................... .................... .................... ....................
F 5165 hCoV-19/USA/WA-S2083/2020|EPI_ISL_497511|2020-06-0 .................... .................... .................... .................... .................... ....................
F 5166 hCoV-19/USA/WA-S2084/2020|EPI_ISL_497512|2020-06-0 .................... .................... .................... .................... .................... ....................
F 5167 hCoV-19/USA/WA-S2085/2020|EPI_ISL_497513|2020-06-0 .................... .................... .................... .................... .................... ....................
F 5168 hCoV-19/USA/WA-S2086/2020|EPI_ISL_497514|2020-06-0 .................... .................... .................... .................... .................... ....................
F 5169 hCoV-19/USA/WA-S2087/2020|EPI_ISL_497515|2020-06-0 .................... .................... .................... .................... .................... ....................
F 5170 hCoV-19/USA/WA-S2088/2020|EPI_ISL_497516|2020-06-0 .................... .................... .................... .................... .................... ....................
F 5171 hCoV-19/USA/WA-S2089/2020|EPI_ISL_497517|2020-06-0 .................... .................... .................... .................... .................... ....................
F 5172 hCoV-19/USA/WA-S2090/2020|EPI_ISL_497518|2020-06-0 .................... .................... .................... .................... .................... ....................
F 5173 hCoV-19/USA/WA-S2091/2020|EPI_ISL_497519|2020-06-0 .................... .................... .................... .................... .................... ....................
F 5174 hCoV-19/USA/WA-S2092/2020|EPI_ISL_497520|2020-06-0 .................... .................... .................... .................... .................... ....................
```

| | | | | | | | |
|---|---|---|---|---|---|---|---|
| F | 5175 hCoV-19/USA/WA-S2093/2020\|EPI_ISL_497521\|2020-06-0 | ···················· | ···················· | ····················· | ···················· | ···················· | ···················· |
| F | 5176 hCoV-19/USA/WA-S2094/2020\|EPI_ISL_497522\|2020-06-0 | ···················· | ···················· | ····················· | ···················· | ···················· | ···················· |
| F | 5177 hCoV-19/USA/WA-S2095/2020\|EPI_ISL_497523\|2020-06-0 | ···················· | ···················· | ····················· | ···················· | ···················· | ···················· |
| F | 5178 hCoV-19/USA/WA-S2096/2020\|EPI_ISL_497524\|2020-06-0 | ···················· | ···················· | ····················· | ···················· | ···················· | ···················· |
| F | 5179 hCoV-19/USA/WA-S2097/2020\|EPI_ISL_497525\|2020-06-0 | ···················· | ···················· | ····················· | ···················· | ···················· | ···················· |
| F | 5180 hCoV-19/USA/WA-S2098/2020\|EPI_ISL_497526\|2020-06-0 | ···················· | ···················· | ····················· | ···················· | ···················· | ···················· |
| F | 5181 hCoV-19/USA/WA-S2099/2020\|EPI_ISL_497527\|2020-06-0 | ···················· | ···················· | ····················· | ···················· | ···················· | ···················· |
| F | 5182 hCoV-19/USA/WA-S2100/2020\|EPI_ISL_497528\|2020-06-0 | ···················· | ···················· | ····················· | ···················· | ···················· | ···················· |
| F | 5183 hCoV-19/USA/WA-S2101/2020\|EPI_ISL_497529\|2020-06-0 | ···················· | ···················· | ····················· | ···················· | ···················· | ···················· |
| F | 5184 hCoV-19/USA/WA-S2102/2020\|EPI_ISL_497530\|2020-06-0 | ···················· | ···················· | ····················· | ···················· | ···················· | ···················· |
| F | 5185 hCoV-19/USA/WA-S2103/2020\|EPI_ISL_497531\|2020-06-0 | ···················· | ···················· | ····················· | ···················· | ···················· | ···················· |
| F | 5186 hCoV-19/USA/WA-S2104/2020\|EPI_ISL_497532\|2020-06-0 | ···················· | ···················· | ····················· | ···················· | ···················· | ···················· |
| F | 5187 hCoV-19/USA/WA-S2105/2020\|EPI_ISL_497533\|2020-06-0 | ···················· | ···················· | ····················· | ···················· | ···················· | ···················· |
| F | 5188 hCoV-19/USA/WA-S2106/2020\|EPI_ISL_497534\|2020-06-0 | ···················· | ···················· | ····················· | ···················· | ···················· | ···················· |
| F | 5189 hCoV-19/USA/WA-S2108/2020\|EPI_ISL_497536\|2020-06-0 | ···················· | ···················· | ····················· | ···················· | ···················· | ···················· |
| F | 5190 hCoV-19/USA/WA-S2109/2020\|EPI_ISL_497537\|2020-06-0 | ···················· | ···················· | ····················· | ···················· | ···················· | ···················· |
| F | 5191 hCoV-19/USA/WA-S2110/2020\|EPI_ISL_497538\|2020-06-0 | ···················· | ···················· | ····················· | ···················· | ···················· | ···················· |
| F | 5192 hCoV-19/USA/WA-S2111/2020\|EPI_ISL_497539\|2020-06-0 | ···················· | ···················· | ····················· | ···················· | ···················· | ···················· |
| F | 5193 hCoV-19/USA/WA-S2112/2020\|EPI_ISL_497540\|2020-06-0 | ···················· | ···················· | ····················· | ···················· | ···················· | ···················· |
| F | 5194 hCoV-19/USA/WA-S2113/2020\|EPI_ISL_497541\|2020-06-0 | ···················· | ···················· | ····················· | ···················· | ···················· | ···················· |
| F | 5195 hCoV-19/USA/WA-S2114/2020\|EPI_ISL_497542\|2020-06-0 | ···················· | ···················· | ····················· | ···················· | ···················· | ···················· |
| F | 5196 hCoV-19/USA/WA-S2116/2020\|EPI_ISL_497544\|2020-06-0 | ···················· | ···················· | ····················· | ···················· | ···················· | ···················· |
| F | 5197 hCoV-19/USA/WA-S2117/2020\|EPI_ISL_497545\|2020-06-0 | ···················· | ···················· | ····················· | ···················· | ···················· | ···················· |
| F | 5198 hCoV-19/USA/WA-S2118/2020\|EPI_ISL_497546\|2020-06-0 | ···················· | ···················· | ····················· | ···················· | ···················· | ···················· |
| F | 5199 hCoV-19/USA/WA-S2119/2020\|EPI_ISL_497547\|2020-06-0 | ···················· | ···················· | ····················· | ···················· | ···················· | ···················· |
| F | 5200 hCoV-19/USA/WA-S2120/2020\|EPI_ISL_497548\|2020-06-0 | ···················· | ···················· | ····················· | ···················· | ···················· | ···················· |
| F | 5201 hCoV-19/USA/WA-S2121/2020\|EPI_ISL_497549\|2020-06-0 | ···················· | ···················· | ····················· | ···················· | ···················· | ···················· |
| F | 5202 hCoV-19/USA/WA-S2122/2020\|EPI_ISL_497550\|2020-06-0 | ···················· | ···················· | ····················· | ···················· | ···················· | ···················· |
| F | 5203 hCoV-19/USA/WA-S2123/2020\|EPI_ISL_497551\|2020-06-0 | ···················· | ···················· | ····················· | ···················· | ···················· | ···················· |
| F | 5204 hCoV-19/USA/WA-S2124/2020\|EPI_ISL_497552\|2020-06-0 | ···················· | ···················· | ····················· | ···················· | ···················· | ···················· |
| F | 5205 hCoV-19/USA/WA-S2125/2020\|EPI_ISL_497553\|2020-06-0 | ···················· | ···················· | ····················· | ···················· | ···················· | ···················· |
| F | 5206 hCoV-19/USA/WA-S2126/2020\|EPI_ISL_497554\|2020-06-0 | ···················· | ···················· | ····················· | ···················· | ···················· | ···················· |
| F | 5207 hCoV-19/USA/WA-S2127/2020\|EPI_ISL_497555\|2020-06-0 | ···················· | ···················· | ····················· | ···················· | ···················· | ···················· |
| F | 5208 hCoV-19/USA/WA-S2128/2020\|EPI_ISL_497556\|2020-06-0 | ···················· | ···················· | ····················· | ···················· | ···················· | ···················· |
| F | 5209 hCoV-19/USA/WA-S2129/2020\|EPI_ISL_497557\|2020-06-0 | ···················· | ···················· | ····················· | ···················· | ···················· | ···················· |
| F | 5210 hCoV-19/USA/WA-S2130/2020\|EPI_ISL_497558\|2020-06-0 | ···················· | ···················· | ····················· | ···················· | ···················· | ···················· |
| F | 5211 hCoV-19/USA/WA-S2131/2020\|EPI_ISL_497559\|2020-06-0 | ···················· | ···················· | ····················· | ···················· | ···················· | ···················· |
| F | 5212 hCoV-19/USA/WA-S2133/2020\|EPI_ISL_497561\|2020-06-0 | ···················· | ···················· | ····················· | ···················· | ···················· | ···················· |
| F | 5213 hCoV-19/USA/WA-S2134/2020\|EPI_ISL_497562\|2020-06-0 | ···················· | ···················· | ····················· | ···················· | ···················· | ···················· |
| F | 5214 hCoV-19/USA/WA-S2136/2020\|EPI_ISL_497564\|2020-06-0 | ···················· | ···················· | ····················· | ···················· | ···················· | ···················· |
| F | 5215 hCoV-19/Bangladesh/BCSIR-NILMRC-275/2020\|EPI_ISL_4 | ···················· | ···················· | ····················· | ···················· | ···················· | ···················· |
| F | 5216 hCoV-19/Bangladesh/BCSIR-NILMRC-281/2020\|EPI_ISL_4 | ···················· | ···················· | ····················· | ···················· | ···················· | ···················· |
| F | 5217 hCoV-19/Bangladesh/BCSIR-NILMRC-283/2020\|EPI_ISL_4 | ···················· | ···················· | ····················· | ···················· | ···················· | ···················· |
| F | 5218 hCoV-19/Bangladesh/BCSIR-NILMRC-288/2020\|EPI_ISL_4 | ···················· | ·A·················· | ····················· | ···················· | ···················· | ··A················· |
| F | 5219 hCoV-19/Bangladesh/BCSIR-NILMRC-296/2020\|EPI_ISL_4 | ···················· | ···················· | ····················· | ···················· | ···················· | ···················· |
| F | 5220 hCoV-19/Bangladesh/BCSIR-NILMRC-305/2020\|EPI_ISL_4 | ···················· | ···················· | ····················· | ···················· | ···················· | ···················· |
| F | 5221 hCoV-19/Bangladesh/BCSIR-NILMRC-311/2020\|EPI_ISL_4 | ···················· | ···················· | ····················· | ···················· | ···················· | ···················· |
| F | 5222 hCoV-19/Bangladesh/BCSIR-NILMRC-312/2020\|EPI_ISL_4 | ···················· | ···················· | ····················· | ···················· | ···················· | ···················· |
| F | 5223 hCoV-19/USA/WA-UW-9368/2020\|EPI_ISL_501160\|2020-05 | ···················· | ···················· | ····················· | ···················· | ···················· | ···················· |
| F | 5224 hCoV-19/USA/WA-UW-9551/2020\|EPI_ISL_501161\|2020-05 | ···················· | ···················· | ····················· | ···················· | ···················· | ···················· |
| F | 5225 hCoV-19/USA/WA-UW-9755/2020\|EPI_ISL_501162\|2020-05 | ···················· | ···················· | ····················· | ···················· | ···················· | ···················· |
| F | 5226 hCoV-19/USA/WA-UW-9798/2020\|EPI_ISL_501163\|2020-05 | ···················· | ···················· | ····················· | ···················· | ···················· | ···················· |
| F | 5227 hCoV-19/USA/WA-UW-9819/2020\|EPI_ISL_501164\|2020-05 | ···················· | ···················· | ····················· | ···················· | ···················· | ···················· |
| F | 5228 hCoV-19/USA/WI-UW-MAYO9/2020\|EPI_ISL_501165\|2020-0 | ···················· | ···················· | ····················· | ···················· | ···················· | ···················· |
| F | 5229 hCoV-19/USA/TX-GCID-192000051B/2020\|EPI_ISL_501167 | ···················· | ···················· | ····················· | ···················· | ···················· | ···················· |
| F | 5230 hCoV-19/USA/TX-GCID-192000207/2020\|EPI_ISL_501169\| | ···················· | ···················· | ····················· | ···················· | ···················· | ···················· |
| F | 5231 hCoV-19/Malaysia/0931/2020\|EPI_ISL_501180\|2020-03- | ···················· | ···················· | ····················· | ···················· | ···················· | ···················· |
| F | 5232 hCoV-19/Malaysia/0956/2020\|EPI_ISL_501181\|2020-03- | ···················· | ···················· | ····················· | ···················· | ···················· | ···················· |
| F | 5233 hCoV-19/Malaysia/1121/2020\|EPI_ISL_501182\|2020-03- | ···················· | ···················· | ····················· | ···················· | ···················· | ···················· |
| F | 5234 hCoV-19/Malaysia/1204/2020\|EPI_ISL_501183\|2020-03- | ···················· | ···················· | ····················· | ···················· | ···················· | ···················· |
| F | 5235 hCoV-19/Malaysia/1399/2020\|EPI_ISL_501184\|2020-03- | ···················· | ···················· | ····················· | ···················· | ···················· | ···················· |
| F | 5236 hCoV-19/Malaysia/1497/2020\|EPI_ISL_501185\|2020-03- | ···················· | ···················· | ····················· | ···················· | ···················· | ···················· |
| F | 5237 hCoV-19/Malaysia/1713/2020\|EPI_ISL_501186\|2020-03- | ···················· | ···················· | ····················· | ···················· | ···················· | ···················· |
| F | 5238 hCoV-19/Malaysia/1798/2020\|EPI_ISL_501187\|2020-03- | ···················· | ···················· | ····················· | ···················· | ···················· | ···················· |
| F | 5239 hCoV-19/Malaysia/2063/2020\|EPI_ISL_501188\|2020-03- | ···················· | ···················· | ····················· | ···················· | ···················· | ···················· |
| F | 5240 hCoV-19/Malaysia/2065/2020\|EPI_ISL_501189\|2020-03- | ···················· | ···················· | ····················· | ···················· | ···················· | ···················· |
| F | 5241 hCoV-19/Malaysia/2079/2020\|EPI_ISL_501190\|2020-03- | ···················· | ···················· | ····················· | ···················· | ···················· | ···················· |
| F | 5242 hCoV-19/Malaysia/2101/2020\|EPI_ISL_501191\|2020-04- | ···················· | ···················· | ····················· | ···················· | ···················· | ···················· |
| F | 5243 hCoV-19/Malaysia/2251/2020\|EPI_ISL_501192\|2020-04- | ···················· | ···················· | ····················· | ···················· | ···················· | ···················· |
| F | 5244 hCoV-19/Malaysia/2297/2020\|EPI_ISL_501193\|2020-03- | ···················· | ···················· | ····················· | ···················· | ···················· | ···················· |
| F | 5245 hCoV-19/Malaysia/2363/2020\|EPI_ISL_501194\|2020-03- | ···················· | ···················· | ····················· | ···················· | ···················· | ···················· |
| F | 5246 hCoV-19/Malaysia/2735/2020\|EPI_ISL_501195\|2020-03- | ···················· | ···················· | ····················· | ···················· | ···················· | ···················· |
| F | 5247 hCoV-19/Malaysia/2811/2020\|EPI_ISL_501196\|2020-03- | ···················· | ···················· | ····················· | ···················· | ···················· | ···················· |
| F | 5248 hCoV-19/Malaysia/2813/2020\|EPI_ISL_501197\|2020-03- | ···················· | ···················· | ····················· | ···················· | ···················· | ···················· |
| F | 5249 hCoV-19/Malaysia/2875/2020\|EPI_ISL_501198\|2020-03- | ···················· | ···················· | ····················· | ···················· | ···················· | ···················· |
| F | 5250 hCoV-19/Malaysia/3012/2020\|EPI_ISL_501199\|2020-03- | ···················· | ···················· | ····················· | ···················· | ···················· | ···················· |
| F | 5251 hCoV-19/Australia/SAP472/2020\|EPI_ISL_508605\|2020- | ···················· | ···················· | ····················· | ···················· | ···················· | ···················· |
| F | 5252 hCoV-19/Australia/SAP473/2020\|EPI_ISL_508606\|2020- | ···················· | ···················· | ····················· | ···················· | ···················· | ···················· |
| F | 5253 hCoV-19/Australia/SAP474/2020\|EPI_ISL_508607\|2020- | ···················· | ···················· | ····················· | ···················· | ···················· | ···················· |
| F | 5254 hCoV-19/USA/WI-UW-898/2020\|EPI_ISL_509900\|2020-07- | ···················· | ···················· | ····················· | ···················· | ···················· | ···················· |

| | | | | | | | | |
|---|---|---|---|---|---|---|---|---|
| F | 5255 | hCoV-19/USA/WI-UW-899/2020\|EPI_ISL_509901\|2020-07- | ···················· | ···················· | ···················· | ···················· | ···················· | ···················· |
| F | 5256 | hCoV-19/USA/WI-UW-900/2020\|EPI_ISL_509902\|2020-07- | ···················· | ···················· | ···················· | ···················· | ···················· | ···················· |
| F | 5257 | hCoV-19/USA/WI-UW-902/2020\|EPI_ISL_509903\|2020-07- | ···················· | ···················· | ···················· | ···················· | ···················· | ···················· |
| F | 5258 | hCoV-19/USA/WI-UW-903/2020\|EPI_ISL_509904\|2020-07- | ···················· | ···················· | ···················· | ···················· | ···················· | ···················· |
| F | 5259 | hCoV-19/USA/WI-UW-904/2020\|EPI_ISL_509905\|2020-07- | ···················· | ···················· | ···················· | ···················· | ···················· | ···················· |
| F | 5260 | hCoV-19/USA/WI-UW-905/2020\|EPI_ISL_509906\|2020-07- | ···················· | ···················· | ···················· | ···················· | ···················· | ···················· |
| F | 5261 | hCoV-19/USA/WI-UW-906/2020\|EPI_ISL_509907\|2020-07- | ···················· | ···················· | ···················· | ···················· | ···················· | ···················· |
| F | 5262 | hCoV-19/USA/WI-UW-907/2020\|EPI_ISL_509908\|2020-07- | ···················· | ···················· | ···················· | ···················· | ···················· | ···················· |
| F | 5263 | hCoV-19/USA/WI-UW-908/2020\|EPI_ISL_509909\|2020-07- | ···················· | ···················· | ···················· | ···················· | ···················· | ···················· |
| F | 5264 | hCoV-19/USA/WI-UW-909/2020\|EPI_ISL_509910\|2020-07- | ···················· | ···················· | ···················· | ···················· | ···················· | ···················· |
| F | 5265 | hCoV-19/USA/WI-UW-910/2020\|EPI_ISL_509911\|2020-07- | ···················· | ···················· | ···················· | ···················· | ···················· | ···················· |
| F | 5266 | hCoV-19/USA/WI-UW-911/2020\|EPI_ISL_509912\|2020-07- | ···················· | ···················· | ···················· | ···················· | ···················· | ···················· |
| F | 5267 | hCoV-19/USA/WI-UW-913/2020\|EPI_ISL_509914\|2020-07- | ···················· | ···················· | ···················· | ···················· | ···················· | ···················· |
| F | 5268 | hCoV-19/USA/WI-UW-914/2020\|EPI_ISL_509915\|2020-07- | ···················· | ···················· | ···················· | ···················· | ···················· | ···················· |
| F | 5269 | hCoV-19/USA/WI-UW-915/2020\|EPI_ISL_509916\|2020-07- | ···················· | ···················· | ···················· | ···················· | ···················· | ···················· |
| F | 5270 | hCoV-19/USA/WI-UW-916/2020\|EPI_ISL_509917\|2020-07- | ···················· | ···················· | ···················· | ···················· | ···················· | ···················· |
| F | 5271 | hCoV-19/USA/WI-UW-917/2020\|EPI_ISL_509918\|2020-07- | ···················· | ···················· | ···················· | ···················· | ···················· | ···················· |
| F | 5272 | hCoV-19/USA/WI-UW-918/2020\|EPI_ISL_509919\|2020-07- | ···················· | ···················· | ···················· | ···················· | ···················· | ···················· |
| F | 5273 | hCoV-19/USA/WI-UW-919/2020\|EPI_ISL_509920\|2020-07- | ···················· | ·A·················· | ···················· | ···················· | ···················· | ···················· |
| F | 5274 | hCoV-19/USA/WI-UW-920/2020\|EPI_ISL_509921\|2020-07- | ···················· | ···················· | ···················· | ···················· | ···················· | ···················· |
| F | 5275 | hCoV-19/USA/WI-UW-922/2020\|EPI_ISL_509922\|2020-07- | ···················· | ···················· | ···················· | ···················· | ···················· | ···················· |
| F | 5276 | hCoV-19/USA/WI-UW-923/2020\|EPI_ISL_509923\|2020-07- | ···················· | ·A·················· | ···················· | ···················· | ···················· | ···················· |
| F | 5277 | hCoV-19/USA/WI-UW-924/2020\|EPI_ISL_509924\|2020-07- | ···················· | ···················· | ···················· | ···················· | ···················· | ···················· |
| F | 5278 | hCoV-19/USA/WI-UW-925/2020\|EPI_ISL_509925\|2020-07- | ···················· | ···················· | ···················· | ···················· | ···················· | ···················· |
| F | 5279 | hCoV-19/USA/WI-UW-926/2020\|EPI_ISL_509926\|2020-07- | ···················· | ···················· | ···················· | ···················· | ···················· | ···················· |
| F | 5280 | hCoV-19/USA/WI-UW-927/2020\|EPI_ISL_509927\|2020-07- | ···················· | ···················· | ···················· | ···················· | ···················· | ···················· |
| F | 5281 | hCoV-19/USA/WI-UW-928/2020\|EPI_ISL_509928\|2020-07- | ···················· | ···················· | ···················· | ···················· | ···················· | ···················· |
| F | 5282 | hCoV-19/USA/WI-UW-929/2020\|EPI_ISL_509929\|2020-07- | ···················· | ···················· | ···················· | ···················· | ···················· | ···················· |
| F | 5284 | hCoV-19/USA/WI-UW-931/2020\|EPI_ISL_509931\|2020-07- | ···················· | ···················· | ···················· | ···················· | ···················· | ···················· |
| F | 5285 | hCoV-19/USA/WI-UW-932/2020\|EPI_ISL_509932\|2020-07- | ···················· | ···················· | ···················· | ···················· | ···················· | ···················· |
| F | 5286 | hCoV-19/USA/WI-UW-933/2020\|EPI_ISL_509933\|2020-05- | ···················· | ···················· | ···················· | ···················· | ···················· | ···················· |
| F | 5287 | hCoV-19/USA/WI-UW-934/2020\|EPI_ISL_509934\|2020-05- | ···················· | ···················· | ···················· | ···················· | ···················· | ···················· |
| F | 5288 | hCoV-19/USA/WI-UW-935/2020\|EPI_ISL_509935\|2020-05- | ···················· | ···················· | ···················· | ···················· | ···················· | ···················· |
| F | 5289 | hCoV-19/USA/WI-UW-936/2020\|EPI_ISL_509936\|2020-05- | ···················· | ···················· | ···················· | ···················· | ···················· | ···················· |
| F | 5290 | hCoV-19/USA/WI-UW-939/2020\|EPI_ISL_509939\|2020-05- | ···················· | ···················· | ···················· | ···················· | ···················· | ···················· |
| F | 5291 | hCoV-19/Portugal/PT0639/2020\|EPI_ISL_510900\|2020-0 | ···················· | ···················· | ···················· | ···················· | ···················· | ···················· |
| F | 5292 | hCoV-19/Portugal/PT0640/2020\|EPI_ISL_510901\|2020-0 | ···················· | ···················· | ···················· | ···················· | ···················· | ···················· |
| F | 5293 | hCoV-19/Portugal/PT0641/2020\|EPI_ISL_510902\|2020-0 | ···················· | ···················· | ···················· | ···················· | ···················· | ···················· |
| F | 5294 | hCoV-19/Portugal/PT0642/2020\|EPI_ISL_510903\|2020-0 | ···················· | ···················· | ···················· | ···················· | ···················· | ···················· |
| F | 5295 | hCoV-19/Portugal/PT0643/2020\|EPI_ISL_510904\|2020-0 | ···················· | ···················· | ···················· | ···················· | ···················· | ···················· |
| F | 5296 | hCoV-19/Portugal/PT0644/2020\|EPI_ISL_510905\|2020-0 | ···················· | ···················· | ···················· | ···················· | ···················· | ···················· |
| F | 5297 | hCoV-19/Portugal/PT0645/2020\|EPI_ISL_510906\|2020-0 | ···················· | ···················· | ···················· | ···················· | ···················· | ···················· |
| F | 5298 | hCoV-19/Portugal/PT0646/2020\|EPI_ISL_510907\|2020-0 | ···················· | ···················· | ···················· | ···················· | ···················· | ···················· |
| F | 5299 | hCoV-19/Portugal/PT0648/2020\|EPI_ISL_510909\|2020-0 | ···················· | ···················· | ···················· | ···················· | ···················· | ···················· |
| F | 5300 | hCoV-19/Portugal/PT0651/2020\|EPI_ISL_510912\|2020-0 | ···················· | ···················· | ···················· | ···················· | ···················· | ···················· |
| F | 5301 | hCoV-19/Portugal/PT0654/2020\|EPI_ISL_510915\|2020-0 | ···················· | ···················· | ···················· | ···················· | ···················· | ···················· |
| F | 5302 | hCoV-19/Portugal/PT0655/2020\|EPI_ISL_510916\|2020-0 | ···················· | ···················· | ···················· | ···················· | ···················· | ···················· |
| F | 5303 | hCoV-19/SaudiArabia/KAUST-MADINAH399/2020\|EPI_ISL_ | ···················· | ···················· | ···················· | ···················· | ···················· | ···················· |
| F | 5304 | hCoV-19/SaudiArabia/KAUST-MADINAH450/2020\|EPI_ISL_ | ···················· | ···················· | ···················· | ···················· | ···················· | ···················· |
| F | 5305 | hCoV-19/SaudiArabia/KAUST-MADINAH452/2020\|EPI_ISL_ | ···················· | ···················· | ···················· | ···················· | ···················· | ···················· |
| F | 5306 | hCoV-19/SaudiArabia/KAUST-MADINAH511/2020\|EPI_ISL_ | ···················· | ···················· | ···················· | ···················· | ···················· | ···················· |
| F | 5307 | hCoV-19/SaudiArabia/KAUST-MADINAH513/2020\|EPI_ISL_ | ···················· | ···················· | ···················· | ···················· | ···················· | ···A················ |
| F | 5308 | hCoV-19/SaudiArabia/KAUST-MADINAH515/2020\|EPI_ISL_ | ···················· | ···················· | ···················· | ···················· | ···················· | ···················· |
| F | 5309 | hCoV-19/SaudiArabia/KAUST-MADINAH519/2020\|EPI_ISL_ | ···················· | ···················· | ···················· | ···················· | ···················· | ···················· |
| F | 5310 | hCoV-19/SaudiArabia/KAUST-MADINAH521/2020\|EPI_ISL_ | ···················· | ···················· | ···················· | ···················· | ···················· | ···················· |
| F | 5311 | hCoV-19/SaudiArabia/KAUST-MADINAH599/2020\|EPI_ISL_ | ···················· | ·A·················· | ···················· | ···················· | ···················· | ···A················ |
| F | 5312 | hCoV-19/SaudiArabia/KAUST-MADINAH603/2020\|EPI_ISL_ | ···················· | ···················· | ···················· | ···················· | ···················· | ···················· |
| F | 5313 | hCoV-19/SaudiArabia/KAUST-MADINAH605/2020\|EPI_ISL_ | ···················· | ···················· | ···················· | ···················· | ···················· | ···················· |
| F | 5314 | hCoV-19/SaudiArabia/KAUST-MADINAH606/2020\|EPI_ISL_ | ···················· | ···················· | ···················· | ···················· | ···················· | ···················· |
| F | 5315 | hCoV-19/SaudiArabia/KAUST-MADINAH609/2020\|EPI_ISL_ | ···················· | ···················· | ···················· | ···················· | ···················· | ···················· |
| F | 5316 | hCoV-19/SaudiArabia/KAUST-MADINAH644/2020\|EPI_ISL_ | ···················· | ···················· | ···················· | ···················· | ···················· | ···················· |
| F | 5317 | hCoV-19/SaudiArabia/KAUST-MADINAH647/2020\|EPI_ISL_ | ···················· | ···················· | ···················· | ···················· | ···················· | ···················· |
| F | 5318 | hCoV-19/SaudiArabia/KAUST-MADINAH655/2020\|EPI_ISL_ | ···················· | ···················· | ···················· | ···················· | ···················· | ···················· |
| F | 5319 | hCoV-19/SaudiArabia/KAUST-MADINAH684/2020\|EPI_ISL_ | ···················· | ···················· | ···················· | ···················· | ···················· | ···················· |
| F | 5320 | hCoV-19/SaudiArabia/KAUST-MADINAH687/2020\|EPI_ISL_ | ···················· | ···················· | ···················· | ···················· | ···················· | ···················· |
| F | 5321 | hCoV-19/SaudiArabia/KAUST-MADINAH688/2020\|EPI_ISL_ | ···················· | ···················· | ···················· | ···················· | ···················· | ···················· |
| F | 5323 | hCoV-19/SaudiArabia/KAUST-MADINAH694/2020\|EPI_ISL_ | ···················· | ···················· | ···················· | ···················· | ···················· | ···················· |
| F | 5324 | hCoV-19/SaudiArabia/KAUST-MADINAH702/2020\|EPI_ISL_ | ···················· | ···················· | ···················· | ···················· | ···················· | ···················· |
| F | 5325 | hCoV-19/SaudiArabia/KAUST-MADINAH704/2020\|EPI_ISL_ | ···················· | ···················· | ···················· | ···················· | ···················· | ···················· |
| F | 5326 | hCoV-19/SaudiArabia/KAUST-MADINAH710/2020\|EPI_ISL_ | ···················· | ···················· | ···················· | ···················· | ···················· | ···················· |
| F | 5327 | hCoV-19/SaudiArabia/KAUST-MADINAH711/2020\|EPI_ISL_ | ···················· | ···················· | ···················· | ···················· | ···················· | ···················· |
| F | 5328 | hCoV-19/SaudiArabia/KAUST-MADINAH712/2020\|EPI_ISL_ | ···················· | ···················· | ···················· | ···················· | ···················· | ···················· |
| F | 5329 | hCoV-19/SaudiArabia/KAUST-MADINAH713/2020\|EPI_ISL_ | ···················· | ···················· | ···················· | ···················· | ···················· | ···················· |
| F | 5330 | hCoV-19/SaudiArabia/KAUST-MADINAH715/2020\|EPI_ISL_ | ···················· | ···················· | ···················· | ···················· | ···················· | ···················· |
| F | 5331 | hCoV-19/SaudiArabia/KAUST-MADINAH716/2020\|EPI_ISL_ | ···················· | ···················· | ···················· | ···················· | ···················· | ···················· |
| F | 5332 | hCoV-19/SaudiArabia/KAUST-MADINAH717/2020\|EPI_ISL_ | ···················· | ···················· | ···················· | ···················· | ···················· | ···················· |
| F | 5333 | hCoV-19/SaudiArabia/KAUST-MADINAH850/2020\|EPI_ISL_ | ···················· | ·A·················· | ···················· | ···················· | ···················· | ···A················ |
| F | 5334 | hCoV-19/SaudiArabia/KAUST-MADINAH851/2020\|EPI_ISL_ | ···················· | ·A·················· | ···················· | ···················· | ···················· | ···A················ |
| F | 5335 | hCoV-19/SaudiArabia/KAUST-MADINAH854/2020\|EPI_ISL_ | ···················· | ···················· | ···················· | ···················· | ···················· | ···················· |
| F | 5336 | hCoV-19/SaudiArabia/KAUST-MADINAH857/2020\|EPI_ISL_ | ···················· | ·A·················· | ···················· | ···················· | ···················· | ···A················ |

| | | | | | | | | | |
|---|---|---|---|---|---|---|---|---|---|
| F | 5337 | hCoV-19/SaudiArabia/KAUST-MADINAH858/2020\|EPI_ISL_ | ···················· | ···················· | ···················· | ···················· | ···················· | ···················· | |
| F | 5338 | hCoV-19/SaudiArabia/KAUST-MADINAH859/2020\|EPI_ISL_ | ···················· | ···················· | ···················· | ···················· | ···················· | ···················· | |
| F | 5339 | hCoV-19/SaudiArabia/KAUST-MADINAH860/2020\|EPI_ISL_ | ···················· | ···················· | ···················· | ···················· | ···················· | ···················· | |
| F | 5340 | hCoV-19/SaudiArabia/KAUST-MADINAH862/2020\|EPI_ISL_ | ···················· | ···················· | ···················· | ···················· | ···················· | ···················· | |
| F | 5341 | hCoV-19/SaudiArabia/KAUST-MADINAH892/2020\|EPI_ISL_ | ···················· | ···················· | ···················· | ···················· | ···················· | ···················· | |
| F | 5342 | hCoV-19/SaudiArabia/KAUST-MADINAH896/2020\|EPI_ISL_ | ···················· | ···················· | ···················· | ···················· | ···················· | ···················· | |
| F | 5343 | hCoV-19/England/PORT-2D1A53/2020\|EPI_ISL_514484\|20 | ···················· | ···················· | ···················· | ···················· | ···················· | ···················· | |
| F | 5344 | hCoV-19/England/PORT-2D267C/2020\|EPI_ISL_514490\|20 | ···················· | ···················· | ···················· | ···················· | ···················· | ···················· | |
| F | 5345 | hCoV-19/South Africa/KRISP-K002324/2020\|EPI_ISL_51 | ···················· | ···················· | ···················· | ···················· | ···················· | ···················· | |
| F | 5346 | hCoV-19/South Africa/KRISP-K002325/2020\|EPI_ISL_51 | ···················· | ···················· | ···················· | ···················· | ···················· | ···················· | |
| F | 5347 | hCoV-19/South Africa/KRISP-K002326/2020\|EPI_ISL_51 | ···················· | ···················· | ···················· | ···················· | ···················· | ···················· | |
| F | 5348 | hCoV-19/South Africa/KRISP-K002327/2020\|EPI_ISL_51 | ···················· | ···················· | ···················· | ···················· | ···················· | ···················· | |
| F | 5349 | hCoV-19/South Africa/KRISP-K002328/2020\|EPI_ISL_51 | ···················· | ···················· | ···················· | ···················· | ···················· | ···················· | |
| F | 5350 | hCoV-19/South Africa/KRISP-K002329/2020\|EPI_ISL_51 | ···················· | ···················· | ···················· | ···················· | ···················· | ···················· | |
| F | 5351 | hCoV-19/South Africa/KRISP-K002331/2020\|EPI_ISL_51 | ···················· | ···················· | ···················· | ···················· | ···················· | ···················· | |
| F | 5352 | hCoV-19/South Africa/KRISP-K002332/2020\|EPI_ISL_51 | ···················· | ···················· | ···················· | ···················· | ···················· | ···················· | |
| F | 5353 | hCoV-19/South Africa/KRISP-K002333/2020\|EPI_ISL_51 | ···················· | ···················· | ···················· | ···················· | ···················· | ···················· | |
| F | 5354 | hCoV-19/South Africa/KRISP-K002335/2020\|EPI_ISL_51 | ···················· | ···················· | ···················· | ···················· | ···················· | ···················· | |
| F | 5355 | hCoV-19/South Africa/KRISP-K002336/2020\|EPI_ISL_51 | ···················· | ···················· | ···················· | ···················· | ···················· | ···················· | |
| F | 5356 | hCoV-19/South Africa/KRISP-K002337/2020\|EPI_ISL_51 | ···················· | ···················· | ···················· | ···················· | ···················· | ···················· | |
| F | 5357 | hCoV-19/South Africa/KRISP-K002338/2020\|EPI_ISL_51 | ···················· | ···················· | ···················· | ···················· | ···················· | ···················· | |
| F | 5358 | hCoV-19/South Africa/KRISP-K002341/2020\|EPI_ISL_51 | ···················· | ···················· | ···················· | ···················· | ···················· | ···················· | |
| F | 5359 | hCoV-19/South Africa/KRISP-K002342/2020\|EPI_ISL_51 | ···················· | ···················· | ···················· | ···················· | ···················· | ···················· | |
| F | 5360 | hCoV-19/South Africa/KRISP-K002343/2020\|EPI_ISL_51 | ···················· | ···················· | ···················· | ···················· | ···················· | ···················· | |
| F | 5361 | hCoV-19/South Africa/KRISP-K002346/2020\|EPI_ISL_51 | ···················· | ···················· | ···················· | ···················· | ···················· | ···················· | |
| F | 5362 | hCoV-19/South Africa/KRISP-K002347/2020\|EPI_ISL_51 | ···················· | ···················· | ···················· | ···················· | ···················· | ···················· | |
| F | 5363 | hCoV-19/South Africa/KRISP-K002348/2020\|EPI_ISL_51 | ···················· | ···················· | ···················· | ···················· | ···················· | ···················· | |
| F | 5364 | hCoV-19/South Africa/KRISP-K002349/2020\|EPI_ISL_51 | ···················· | ···················· | ···················· | ···················· | NNN················· | ···················· | 1 |
| F | 5365 | hCoV-19/South Africa/KRISP-K002350/2020\|EPI_ISL_51 | ···················· | ···················· | ···················· | ···················· | ···················· | ···················· | |
| F | 5366 | hCoV-19/South Africa/KRISP-K002351/2020\|EPI_ISL_51 | ···················· | ···················· | ···················· | ···················· | ···················· | ···················· | |
| F | 5367 | hCoV-19/South Africa/KRISP-K002352/2020\|EPI_ISL_51 | ···················· | ···················· | ···················· | ···················· | ···················· | ···················· | |
| F | 5368 | hCoV-19/South Africa/KRISP-K002353/2020\|EPI_ISL_51 | ···················· | ···················· | ···················· | ···················· | ···················· | ···················· | |
| F | 5369 | hCoV-19/South Africa/KRISP-K002357/2020\|EPI_ISL_51 | ···················· | ···················· | ···················· | ···················· | ···················· | ···················· | |
| F | 5370 | hCoV-19/South Africa/KRISP-K002358/2020\|EPI_ISL_51 | ···················· | ···················· | ···················· | ···················· | ···················· | ···················· | |
| F | 5371 | hCoV-19/South Africa/KRISP-K002359/2020\|EPI_ISL_51 | ···················· | ···················· | ···················· | ···················· | ···················· | ···················· | |
| F | 5372 | hCoV-19/South Africa/KRISP-K002363/2020\|EPI_ISL_51 | ···················· | ···················· | ···················· | ···················· | ···················· | ···················· | |
| F | 5373 | hCoV-19/South Africa/KRISP-K002364/2020\|EPI_ISL_51 | ···················· | ···················· | ···················· | ···················· | ···················· | ···················· | |
| F | 5374 | hCoV-19/South Africa/KRISP-K002365/2020\|EPI_ISL_51 | ···················· | ···················· | ···················· | ···················· | ···················· | ···················· | |
| F | 5375 | hCoV-19/South Africa/KRISP-K002366/2020\|EPI_ISL_51 | ···················· | ···················· | ···················· | ···················· | ···················· | ···················· | |
| F | 5376 | hCoV-19/South Africa/KRISP-K002368/2020\|EPI_ISL_51 | ···················· | ···················· | ···················· | ···················· | ···················· | ···················· | |
| F | 5377 | hCoV-19/Bangladesh/BCSIR-NILMRC-339/2020\|EPI_ISL_4 | ···················· | ···················· | ···················· | ···················· | ···················· | ···················· | |
| F | 5378 | hCoV-19/Bangladesh/BCSIR-NILMRC-340/2020\|EPI_ISL_4 | ···················· | ···················· | ···················· | ···················· | ···················· | ···················· | |
| F | 5379 | hCoV-19/Bangladesh/BCSIR-NILMRC-341/2020\|EPI_ISL_4 | ···················· | ···················· | ···················· | ···················· | ···················· | ···················· | |
| F | 5380 | hCoV-19/Bangladesh/BCSIR-NILMRC-345/2020\|EPI_ISL_4 | ···················· | ···················· | ···················· | ···················· | ···················· | ···················· | |
| F | 5381 | hCoV-19/Bangladesh/BCSIR-NILMRC-346/2020\|EPI_ISL_4 | ···················· | ···················· | ···················· | ···················· | ···················· | ···················· | |
| F | 5382 | hCoV-19/Bangladesh/BCSIR-NILMRC-349/2020\|EPI_ISL_4 | ···················· | ···················· | ···················· | ···················· | ···················· | ···················· | |
| F | 5383 | hCoV-19/Bangladesh/BCSIR-NILMRC-259/2020\|EPI_ISL_4 | ···················· | ···················· | ···················· | ···················· | ···················· | ···················· | |
| F | 5384 | hCoV-19/USA/WA-UW-13332/2020\|EPI_ISL_501140\|2020-0 | ···················· | ···················· | ···················· | ···················· | ···················· | ···················· | |
| F | 5385 | hCoV-19/USA/WA-UW-13356/2020\|EPI_ISL_501141\|2020-0 | ···················· | ···················· | ···················· | ···················· | ···················· | ···················· | |
| F | 5386 | hCoV-19/USA/WA-UW-13357/2020\|EPI_ISL_501142\|2020-0 | ···················· | ···················· | ···················· | ···················· | ···················· | ···················· | |
| F | 5387 | hCoV-19/USA/WA-UW-13364/2020\|EPI_ISL_501143\|2020-0 | ···················· | ···················· | ···················· | ···················· | ···················· | ···················· | |
| F | 5388 | hCoV-19/USA/WA-UW-13366/2020\|EPI_ISL_501144\|2020-0 | ···················· | ·A·················· | ···················· | ···················· | ···················· | ···················· | |
| F | 5389 | hCoV-19/USA/WA-UW-13367/2020\|EPI_ISL_501145\|2020-0 | ···················· | ···················· | ···················· | ···················· | ···················· | ···················· | |
| F | 5390 | hCoV-19/USA/WA-UW-13386/2020\|EPI_ISL_501146\|2020-0 | ···················· | ···················· | ···················· | ···················· | ···················· | ···················· | |
| F | 5391 | hCoV-19/USA/WA-UW-13395/2020\|EPI_ISL_501147\|2020-0 | ···················· | ···················· | ···················· | ···················· | ···················· | ···················· | |
| F | 5392 | hCoV-19/USA/WA-UW-13400/2020\|EPI_ISL_501148\|2020-0 | ···················· | ···················· | ···················· | ···················· | ···················· | ···················· | |
| F | 5393 | hCoV-19/USA/WA-UW-13401/2020\|EPI_ISL_501149\|2020-0 | ···················· | ·A·················· | ···················· | ···················· | ···················· | ···················· | |
| F | 5394 | hCoV-19/USA/WA-UW-13402/2020\|EPI_ISL_501150\|2020-0 | ···················· | ···················· | ···················· | ···················· | ···················· | ···················· | |
| F | 5395 | hCoV-19/USA/WA-UW-7745/2020\|EPI_ISL_501151\|2020-04 | ···················· | ···················· | ···················· | ···················· | ···················· | ···················· | |
| F | 5396 | hCoV-19/USA/WA-UW-7764/2020\|EPI_ISL_501152\|2020-04 | ···················· | ···················· | ···················· | ···················· | ···················· | ···················· | |
| F | 5397 | hCoV-19/USA/WA-UW-7882/2020\|EPI_ISL_501153\|2020-04 | ···················· | ···················· | ···················· | ···················· | ···················· | ···················· | |
| F | 5398 | hCoV-19/USA/WA-UW-8046/2020\|EPI_ISL_501154\|2020-04 | ···················· | ···················· | ···················· | ···················· | ···················· | ···················· | |
| F | 5399 | hCoV-19/USA/WA-UW-8686/2020\|EPI_ISL_501155\|2020-05 | ···················· | ···················· | ···················· | ···················· | ···················· | ···················· | |
| F | 5400 | hCoV-19/USA/WA-UW-8803/2020\|EPI_ISL_501156\|2020-05 | ···················· | ···················· | ···················· | ···················· | ···················· | ···················· | |
| F | 5401 | hCoV-19/USA/WA-UW-8845/2020\|EPI_ISL_501157\|2020-05 | ···················· | ···················· | ···················· | ···················· | ···················· | ···················· | |
| F | 5402 | hCoV-19/USA/WA-UW-9032/2020\|EPI_ISL_501158\|2020-05 | ···················· | ···················· | ···················· | ···················· | ···················· | ···················· | |
| F | 5403 | hCoV-19/USA/WA-UW-9291/2020\|EPI_ISL_501159\|2020-05 | ···················· | ···················· | ···················· | ···················· | ···················· | ···················· | |
| F | 5404 | hCoV-19/USA/TX-GCID-192000254/2020\|EPI_ISL_501171\| | ···················· | ···················· | ···················· | ···················· | ···················· | ···················· | |
| F | 5405 | hCoV-19/USA/TX-GCID-192000296/2020\|EPI_ISL_501172\| | ···················· | ···················· | ···················· | ···················· | ···················· | ···················· | |
| F | 5406 | hCoV-19/USA/TX-GCID-192000331/2020\|EPI_ISL_501173\| | ···················· | ···················· | ···················· | ···················· | ···················· | ···················· | |
| F | 5407 | hCoV-19/Malaysia/0121/2020\|EPI_ISL_501176\|2020-03- | ···················· | ···················· | ···················· | ···················· | ···················· | ···················· | |
| F | 5408 | hCoV-19/Malaysia/0309/2020\|EPI_ISL_501177\|2020-03- | ···················· | ···················· | ···················· | ···················· | ···················· | ···················· | |
| F | 5409 | hCoV-19/Malaysia/0478/2020\|EPI_ISL_501178\|2020-03- | ···················· | ···················· | ···················· | ···················· | ···················· | ···················· | |
| F | 5410 | hCoV-19/Malaysia/0784/2020\|EPI_ISL_501179\|2020-03- | ···················· | ···················· | ···················· | ···················· | ···················· | ···················· | |
| F | 5411 | hCoV-19/Russia/YAR-RII-24773S/2020\|EPI_ISL_507300\| | ···················· | ···················· | ···················· | ···················· | ···················· | ···················· | |
| F | 5421 | hCoV-19/Spain/CT-IBV-005626/2020\|EPI_ISL_508661\|20 | ···················· | ···················· | ···················· | ···················· | ·NN················· | ···················· | |
| F | 5425 | hCoV-19/Germany/BW-UT-009/2020\|EPI_ISL_508688\|2020 | ···················· | ···················· | ···················· | ···················· | ···················· | ···················· | |
| F | 5426 | hCoV-19/USA/WI-UW-940/2020\|EPI_ISL_509940\|2020-05- | ···················· | ···················· | ···················· | ···················· | ···················· | ···················· | |
| F | 5427 | hCoV-19/USA/WI-UW-941/2020\|EPI_ISL_509941\|2020-05- | ···················· | ···················· | ···················· | ···················· | ···················· | ···················· | |
| F | 5428 | hCoV-19/USA/WI-UW-942/2020\|EPI_ISL_509942\|2020-05- | ···················· | ···················· | ···················· | ···················· | ···················· | ···················· | |

| | | | | | | | | |
|---|---|---|---|---|---|---|---|---|
| F | 5430 | hCoV-19/USA/WI-UW-944/2020\|EPI_ISL_509944\|2020-05- | ···················· | ···················· | ····················· | ···················· | ···················· | ···················· |
| F | 5431 | hCoV-19/USA/WI-UW-946/2020\|EPI_ISL_509945\|2020-05- | ···················· | ···················· | ····················· | ···················· | ···················· | ···················· |
| F | 5432 | hCoV-19/USA/WI-UW-947/2020\|EPI_ISL_509946\|2020-05- | ···················· | ···················· | ····················· | ···················· | ···················· | ···················· |
| F | 5433 | hCoV-19/USA/WI-UW-948/2020\|EPI_ISL_509947\|2020-05- | ···················· | ···················· | ····················· | ···················· | ···················· | ···················· |
| F | 5434 | hCoV-19/USA/WI-UW-949/2020\|EPI_ISL_509948\|2020-05- | ···················· | ···················· | ····················· | ···················· | ···················· | ···················· |
| F | 5435 | hCoV-19/USA/WI-UW-950/2020\|EPI_ISL_509949\|2020-05- | ···················· | ···················· | ····················· | ···················· | ···················· | ···················· |
| F | 5436 | hCoV-19/USA/WI-UW-951/2020\|EPI_ISL_509950\|2020-05- | ···················· | ···················· | ····················· | ···················· | ···················· | ···················· |
| F | 5437 | hCoV-19/USA/WI-UW-952/2020\|EPI_ISL_509951\|2020-05- | ···················· | ···················· | ····················· | ···················· | ···················· | ···················· |
| F | 5438 | hCoV-19/USA/WI-UW-953/2020\|EPI_ISL_509952\|2020-05- | ···················· | ···················· | ····················· | ···················· | ···················· | ···················· |
| F | 5439 | hCoV-19/USA/WI-UW-954/2020\|EPI_ISL_509953\|2020-05- | ···················· | ···················· | ····················· | ···················· | ···················· | ···················· |
| F | 5440 | hCoV-19/USA/WI-UW-955/2020\|EPI_ISL_509954\|2020-05- | ···················· | ···················· | ····················· | ···················· | ···················· | ···················· |
| F | 5441 | hCoV-19/USA/WI-UW-956/2020\|EPI_ISL_509955\|2020-05- | ···················· | ···················· | ····················· | ···················· | ···················· | ···················· |
| F | 5442 | hCoV-19/USA/WI-UW-957/2020\|EPI_ISL_509956\|2020-05- | ···················· | ···················· | ····················· | ···················· | ···················· | ···················· |
| F | 5443 | hCoV-19/USA/WI-UW-958/2020\|EPI_ISL_509957\|2020-05- | ···················· | ···················· | ····················· | ···················· | ···················· | ···················· |
| F | 5444 | hCoV-19/USA/WI-UW-959/2020\|EPI_ISL_509958\|2020-05- | ···················· | ···················· | ····················· | ···················· | ···················· | ···················· |
| F | 5445 | hCoV-19/USA/WI-UW-960/2020\|EPI_ISL_509959\|2020-05- | ···················· | ···················· | ····················· | ···················· | ···················· | ···················· |
| F | 5446 | hCoV-19/USA/WI-UW-962/2020\|EPI_ISL_509960\|2020-06- | ···················· | ···················· | ····················· | ···················· | ···················· | ···················· |
| F | 5447 | hCoV-19/USA/WI-UW-963/2020\|EPI_ISL_509961\|2020-06- | ···················· | ···················· | ····················· | ···················· | ···················· | ···················· |
| F | 5448 | hCoV-19/USA/WI-UW-964/2020\|EPI_ISL_509962\|2020-06- | ···················· | ···················· | ····················· | ···················· | ···················· | ···················· |
| F | 5449 | hCoV-19/USA/WI-UW-965/2020\|EPI_ISL_509963\|2020-05- | ···················· | ···················· | ····················· | ···················· | ···················· | ···················· |
| F | 5450 | hCoV-19/USA/WI-UW-966/2020\|EPI_ISL_509964\|2020-07- | ···················· | ···················· | ····················· | ···················· | ···················· | ···················· |
| F | 5451 | hCoV-19/USA/WI-UW-967/2020\|EPI_ISL_509965\|2020-07- | ···················· | ···················· | ····················· | ···················· | ···················· | ···················· |
| F | 5452 | hCoV-19/USA/WI-UW-968/2020\|EPI_ISL_509966\|2020-07- | ···················· | ···················· | ····················· | ···················· | ···················· | ···················· |
| F | 5453 | hCoV-19/USA/WI-UW-969/2020\|EPI_ISL_509967\|2020-07- | ···················· | ···················· | ····················· | ···················· | ···················· | ···················· |
| F | 5454 | hCoV-19/USA/WI-UW-971/2020\|EPI_ISL_509969\|2020-07- | ···················· | ···················· | ····················· | ···················· | ···················· | ···················· |
| F | 5455 | hCoV-19/USA/WI-UW-972/2020\|EPI_ISL_509970\|2020-07- | ···················· | ···················· | ····················· | ···················· | ···················· | ···················· |
| F | 5456 | hCoV-19/USA/WI-UW-973/2020\|EPI_ISL_509971\|2020-07- | ···················· | ···················· | ····················· | ···················· | ···················· | ···················· |
| F | 5457 | hCoV-19/USA/WI-UW-974/2020\|EPI_ISL_509972\|2020-07- | ···················· | ···················· | ····················· | ···················· | ···················· | ···················· |
| F | 5458 | hCoV-19/USA/WI-UW-976/2020\|EPI_ISL_509973\|2020-07- | ···················· | ···················· | ····················· | ···················· | ···················· | ···················· |
| F | 5459 | hCoV-19/USA/WI-UW-977/2020\|EPI_ISL_509974\|2020-07- | ···················· | ···················· | ····················· | ···················· | ···················· | ···················· |
| F | 5460 | hCoV-19/USA/WI-UW-978/2020\|EPI_ISL_509975\|2020-07- | ···················· | ···················· | ····················· | ···················· | ···················· | ···················· |
| F | 5461 | hCoV-19/USA/WI-UW-980/2020\|EPI_ISL_509976\|2020-07- | ···················· | ···················· | ····················· | ···················· | ···················· | ···················· |
| F | 5462 | hCoV-19/USA/WI-UW-981/2020\|EPI_ISL_509977\|2020-07- | ···················· | ···················· | ····················· | ···················· | ···················· | ···················· |
| F | 5463 | hCoV-19/USA/WI-UW-982/2020\|EPI_ISL_509978\|2020-07- | ···················· | ···················· | ····················· | ···················· | ···················· | ···················· |
| F | 5464 | hCoV-19/USA/WI-UW-983/2020\|EPI_ISL_509979\|2020-07- | ···················· | ···················· | ····················· | ···················· | ···················· | ···················· |
| F | 5465 | hCoV-19/USA/WI-UW-984/2020\|EPI_ISL_509980\|2020-07- | ···················· | ···················· | ····················· | ···················· | ···················· | ···················· |
| F | 5466 | hCoV-19/USA/WI-UW-985/2020\|EPI_ISL_509981\|2020-07- | ···················· | ···················· | ····················· | ···················· | ···················· | ···················· |
| F | 5467 | hCoV-19/USA/WI-UW-986/2020\|EPI_ISL_509982\|2020-07- | ···················· | ···················· | ····················· | ···················· | ···················· | ···················· |
| F | 5468 | hCoV-19/USA/WI-UW-988/2020\|EPI_ISL_509983\|2020-07- | ···················· | ···················· | ····················· | ···················· | ···················· | ···················· |
| F | 5469 | hCoV-19/USA/WI-UW-989/2020\|EPI_ISL_509984\|2020-07- | ···················· | ·A·················· | ····················· | ···················· | ···················· | ···················· |
| F | 5470 | hCoV-19/USA/WI-UW-990/2020\|EPI_ISL_509985\|2020-07- | ···················· | ···················· | ····················· | ···················· | ···················· | ···················· |
| F | 5471 | hCoV-19/USA/WI-UW-991/2020\|EPI_ISL_509986\|2020-07- | ···················· | ···················· | ····················· | ···················· | ···················· | ···················· |
| F | 5473 | hCoV-19/USA/WI-UW-993/2020\|EPI_ISL_509988\|2020-07- | ···················· | ···················· | ····················· | ···················· | ···················· | ···················· |
| F | 5474 | hCoV-19/USA/WI-UW-995/2020\|EPI_ISL_509990\|2020-07- | ···················· | ···················· | ····················· | ···················· | ···················· | ···················· |
| F | 5475 | hCoV-19/USA/WI-UW-997/2020\|EPI_ISL_509991\|2020-07- | ···················· | ···················· | ····················· | ···················· | ···················· | ···················· |
| F | 5476 | hCoV-19/USA/WI-UW-998/2020\|EPI_ISL_509992\|2020-07- | ···················· | ···················· | ····················· | ···················· | ···················· | ···················· |
| F | 5477 | hCoV-19/USA/WI-UW-846/2020\|EPI_ISL_509994\|2020-07- | ···················· | ···················· | ····················· | ···················· | ···················· | ···················· |
| F | 5478 | hCoV-19/USA/IL-UW-901/2020\|EPI_ISL_509995\|2020-07- | ···················· | ···················· | ····················· | ···················· | ···················· | ···················· |
| F | 5479 | hCoV-19/USA/WI-UW-811/2020\|EPI_ISL_509996\|2020-06- | ···················· | ···················· | ····················· | ···················· | ···················· | ···················· |
| F | 5480 | hCoV-19/USA/WI-UW-814/2020\|EPI_ISL_509997\|2020-06- | ···················· | ···················· | ····················· | ···················· | ···················· | ···················· |
| F | 5481 | hCoV-19/USA/WI-UW-945/2020\|EPI_ISL_509998\|2020-05- | ···················· | ···················· | ····················· | ···················· | ···················· | ···················· |
| F | 5482 | hCoV-19/Portugal/PT0662/2020\|EPI_ISL_510921\|2020-0 | ···················· | ···················· | ····················· | ···················· | ···················· | ···················· |
| F | 5483 | hCoV-19/Portugal/PT0664/2020\|EPI_ISL_510923\|2020-0 | ···················· | ···················· | ····················· | ···················· | ···················· | ···················· |
| F | 5484 | hCoV-19/Portugal/PT0665/2020\|EPI_ISL_510924\|2020-0 | ···················· | ···················· | ····················· | ···················· | ···················· | ···················· |
| F | 5485 | hCoV-19/Portugal/PT0666/2020\|EPI_ISL_510925\|2020-0 | ···················· | ···················· | ····················· | ···················· | ···················· | ···················· |
| F | 5486 | hCoV-19/Portugal/PT0667/2020\|EPI_ISL_510926\|2020-0 | ···················· | ···················· | ····················· | ···················· | ···················· | ···················· |
| F | 5487 | hCoV-19/Portugal/PT0668/2020\|EPI_ISL_510927\|2020-0 | ···················· | ···················· | ····················· | ···················· | ···················· | ···················· |
| F | 5488 | hCoV-19/Portugal/PT0669/2020\|EPI_ISL_510928\|2020-0 | ···················· | ···················· | ····················· | ···················· | ···················· | ···················· |
| F | 5489 | hCoV-19/Portugal/PT0670/2020\|EPI_ISL_510929\|2020-0 | ···················· | ···················· | ····················· | ···················· | ···················· | ···················· |
| F | 5491 | hCoV-19/Portugal/PT0673/2020\|EPI_ISL_510932\|2020-0 | ···················· | ···················· | ····················· | ···················· | ···················· | ···················· |
| F | 5492 | hCoV-19/Portugal/PT0674/2020\|EPI_ISL_510933\|2020-0 | ···················· | ···················· | ····················· | ···················· | ···················· | ···················· |
| F | 5493 | hCoV-19/Portugal/PT0675/2020\|EPI_ISL_510934\|2020-0 | ···················· | ···················· | ····················· | ···················· | ···················· | ···················· |
| F | 5494 | hCoV-19/Portugal/PT0676/2020\|EPI_ISL_510935\|2020-0 | ···················· | ···················· | ····················· | ···················· | ···················· | ···················· |
| F | 5495 | hCoV-19/Portugal/PT0677/2020\|EPI_ISL_510936\|2020-0 | ···················· | ···················· | ····················· | ···················· | ···················· | ···················· |
| F | 5496 | hCoV-19/Portugal/PT0678/2020\|EPI_ISL_510937\|2020-0 | ···················· | ···················· | ····················· | ···················· | ···················· | ···················· |
| F | 5497 | hCoV-19/Portugal/PT0679/2020\|EPI_ISL_510938\|2020-0 | ···················· | ···················· | ····················· | ···················· | ···················· | ···················· |
| F | 5498 | hCoV-19/Portugal/PT0680/2020\|EPI_ISL_510939\|2020-0 | ···················· | ···················· | ····················· | ···················· | ···················· | ···················· |
| F | 5499 | hCoV-19/Portugal/PT0681/2020\|EPI_ISL_510940\|2020-0 | ···················· | ···················· | ····················· | ···················· | ···················· | ···················· |
| F | 5500 | hCoV-19/Portugal/PT0682/2020\|EPI_ISL_510941\|2020-0 | ···················· | ···················· | ····················· | ···················· | ···················· | ···················· |
| F | 5501 | hCoV-19/Portugal/PT0683/2020\|EPI_ISL_510942\|2020-0 | ···················· | ···················· | ····················· | ···················· | ···················· | ···················· |
| F | 5502 | hCoV-19/Portugal/PT0684/2020\|EPI_ISL_510943\|2020-0 | ···················· | ···················· | ····················· | ···················· | ···················· | ···················· |
| F | 5503 | hCoV-19/Portugal/PT0685/2020\|EPI_ISL_510944\|2020-0 | ···················· | ···················· | ····················· | ···················· | ···················· | ···················· |
| F | 5504 | hCoV-19/Portugal/PT0686/2020\|EPI_ISL_510945\|2020-0 | ···················· | ···················· | ····················· | ···················· | ···················· | ···················· |
| F | 5506 | hCoV-19/Portugal/PT0688/2020\|EPI_ISL_510947\|2020-0 | ···················· | ···················· | ····················· | ···················· | ···················· | ···················· |
| F | 5507 | hCoV-19/Portugal/PT0689/2020\|EPI_ISL_510948\|2020-0 | ···················· | ···················· | ····················· | ···················· | ···················· | ···················· |
| F | 5508 | hCoV-19/Portugal/PT0690/2020\|EPI_ISL_510949\|2020-0 | ···················· | ···················· | ····················· | ···················· | ···················· | ···················· |
| F | 5509 | hCoV-19/Portugal/PT0691/2020\|EPI_ISL_510950\|2020-0 | ···················· | ···················· | ····················· | ···················· | ···················· | ···················· |
| F | 5510 | hCoV-19/Portugal/PT0692/2020\|EPI_ISL_510951\|2020-0 | ···················· | ···················· | ····················· | ···················· | ···················· | ···················· |
| F | 5511 | hCoV-19/Portugal/PT0693/2020\|EPI_ISL_510952\|2020-0 | ···················· | ···················· | ····················· | ···················· | ···················· | ···················· |
| F | 5512 | hCoV-19/Portugal/PT0694/2020\|EPI_ISL_510953\|2020-0 | ···················· | ···················· | ····················· | ···················· | ···················· | ···················· |

| | | | | | | | | |
|---|---|---|---|---|---|---|---|---|
| F | 5513 | hCoV-19/Portugal/PT0695/2020\|EPI_ISL_510954\|2020-0 | ···················· | ···················· | ···················· | ···················· | ···················· | ···················· |
| F | 5514 | hCoV-19/Portugal/PT0697/2020\|EPI_ISL_510956\|2020-0 | ···················· | ···················· | ···················· | ···················· | ···················· | ···················· |
| F | 5515 | hCoV-19/Portugal/PT0699/2020\|EPI_ISL_510958\|2020-0 | ···················· | ···················· | ···················· | ···················· | ···················· | ···················· |
| F | 5516 | hCoV-19/Portugal/PT0700/2020\|EPI_ISL_510959\|2020-0 | ···················· | ···················· | ···················· | ···················· | ···················· | ···················· |
| F | 5517 | hCoV-19/Portugal/PT0701/2020\|EPI_ISL_510960\|2020-0 | ···················· | ···················· | ···················· | ···················· | ···················· | ···················· |
| F | 5518 | hCoV-19/Portugal/PT0702/2020\|EPI_ISL_510961\|2020-0 | ···················· | ···················· | ···················· | ···················· | ···················· | ···················· |
| F | 5519 | hCoV-19/Portugal/PT0703/2020\|EPI_ISL_510962\|2020-0 | ···················· | ···················· | ···················· | ···················· | ···················· | ···················· |
| F | 5520 | hCoV-19/Portugal/PT0704/2020\|EPI_ISL_510963\|2020-0 | ···················· | ···················· | ···················· | ···················· | ···················· | ···················· |
| F | 5521 | hCoV-19/Portugal/PT0705/2020\|EPI_ISL_510964\|2020-0 | ···················· | ···················· | ···················· | ···················· | ···················· | ···················· |
| F | 5522 | hCoV-19/Portugal/PT0706/2020\|EPI_ISL_510965\|2020-0 | ···················· | ···················· | ···················· | ···················· | ···················· | ···················· |
| F | 5523 | hCoV-19/Portugal/PT0707/2020\|EPI_ISL_510966\|2020-0 | ···················· | ···················· | ···················· | ···················· | ···················· | ···················· |
| F | 5524 | hCoV-19/Portugal/PT0708/2020\|EPI_ISL_510967\|2020-0 | ···················· | ···················· | ···················· | ···················· | ···················· | ···················· |
| F | 5525 | hCoV-19/Portugal/PT0709/2020\|EPI_ISL_510968\|2020-0 | ···················· | ···················· | ···················· | ···················· | ···················· | ···················· |
| F | 5526 | hCoV-19/Portugal/PT0710/2020\|EPI_ISL_510969\|2020-0 | ···T················ | ···················· | ···················· | ···················· | ···················· | ···················· |
| F | 5527 | hCoV-19/Portugal/PT0711/2020\|EPI_ISL_510970\|2020-0 | ···················· | ···················· | ···················· | ···················· | ···················· | ···················· |
| F | 5528 | hCoV-19/Portugal/PT0712/2020\|EPI_ISL_510971\|2020-0 | ···················· | ···················· | ···················· | ···················· | ···················· | ···················· |
| F | 5529 | hCoV-19/Portugal/PT0713/2020\|EPI_ISL_510972\|2020-0 | ···················· | ···················· | ···················· | ···················· | ···················· | ···················· |
| F | 5530 | hCoV-19/Portugal/PT0714/2020\|EPI_ISL_510973\|2020-0 | ···················· | ···················· | ···················· | ···················· | ···················· | ···················· |
| F | 5531 | hCoV-19/Portugal/PT0715/2020\|EPI_ISL_510974\|2020-0 | ···················· | ···················· | ···················· | ···················· | ···················· | ···················· |
| F | 5532 | hCoV-19/Portugal/PT0716/2020\|EPI_ISL_510975\|2020-0 | ···················· | ···················· | ···················· | ···················· | ···················· | ···················· |
| F | 5533 | hCoV-19/Portugal/PT0717/2020\|EPI_ISL_510976\|2020-0 | ···················· | ···················· | ···················· | ···················· | ···················· | ···················· |
| F | 5534 | hCoV-19/Portugal/PT0719/2020\|EPI_ISL_510978\|2020-0 | ···················· | ···················· | ···················· | ···················· | ···················· | ···················· |
| F | 5535 | hCoV-19/Portugal/PT0722/2020\|EPI_ISL_510981\|2020-0 | ···················· | ···················· | ···················· | ···················· | ···················· | ···················· |
| F | 5536 | hCoV-19/Portugal/PT0723/2020\|EPI_ISL_510982\|2020-0 | ···················· | ···················· | ···················· | ···················· | ···················· | ···················· |
| F | 5537 | hCoV-19/Portugal/PT0724/2020\|EPI_ISL_510983\|2020-0 | ···················· | ···················· | ···················· | ···················· | ···················· | ···················· |
| F | 5538 | hCoV-19/Portugal/PT0725/2020\|EPI_ISL_510984\|2020-0 | ···················· | ···················· | ···················· | ···················· | ···················· | ···················· |
| F | 5539 | hCoV-19/Portugal/PT0728/2020\|EPI_ISL_510987\|2020-0 | ···················· | ···················· | ···················· | ···················· | ···················· | ···················· |
| F | 5540 | hCoV-19/Portugal/PT0732/2020\|EPI_ISL_510991\|2020-0 | ···················· | ···················· | ···················· | ···················· | ···················· | ···················· |
| F | 5541 | hCoV-19/Portugal/PT0733/2020\|EPI_ISL_510992\|2020-0 | ···················· | ···················· | ···················· | ···················· | ···················· | ···················· |
| F | 5542 | hCoV-19/Portugal/PT0735/2020\|EPI_ISL_510994\|2020-0 | ···················· | ···················· | ···················· | ···················· | ···················· | ···················· |
| F | 5543 | hCoV-19/Portugal/PT0736/2020\|EPI_ISL_510995\|2020-0 | ···················· | ···················· | ···················· | ···················· | ···················· | ···················· |
| F | 5544 | hCoV-19/Portugal/PT0738/2020\|EPI_ISL_510997\|2020-0 | ···················· | ···················· | ···················· | ···················· | ···················· | ···················· |
| F | 5545 | hCoV-19/Portugal/PT0740/2020\|EPI_ISL_510999\|2020-0 | ···················· | ···················· | ···················· | ···················· | ···················· | ···················· |
| F | 5546 | hCoV-19/Portugal/PT1244/2020\|EPI_ISL_511205\|2020-0 | ···················· | ···················· | ···················· | ···················· | ···················· | ···················· |
| F | 5547 | hCoV-19/Portugal/PT1245/2020\|EPI_ISL_511206\|2020-0 | ···················· | ···················· | ···················· | ···················· | ···················· | ···················· |
| F | 5548 | hCoV-19/Portugal/PT1248/2020\|EPI_ISL_511209\|2020-0 | ···················· | ···················· | ···················· | ···················· | ···················· | ···················· |
| F | 5549 | hCoV-19/Wales/PHWC-169936/2020\|EPI_ISL_512536\|2020 | ···················· | ···················· | ···················· | ···················· | ···················· | ···················· |
| F | 5550 | hCoV-19/Wales/PHWC-169945/2020\|EPI_ISL_512537\|2020 | ···················· | ···················· | ···················· | ···················· | ···················· | ···················· |
| F | 5551 | hCoV-19/Wales/PHWC-169963/2020\|EPI_ISL_512539\|2020 | ···················· | ···················· | ···················· | ···················· | ···················· | ···················· |
| F | 5552 | hCoV-19/SaudiArabia/KAUST-MADINAH1115/2020\|EPI_ISL | ···················· | ·A·················· | ···················· | ···················· | ···················· | ···A················ |
| F | 5553 | hCoV-19/SaudiArabia/KAUST-MADINAH1116/2020\|EPI_ISL | ···················· | ·A·················· | ···················· | ···················· | ···················· | ···A················ |
| F | 5554 | hCoV-19/SaudiArabia/KAUST-MADINAH273/2020\|EPI_ISL_ | ···················· | ···················· | ···················· | ···················· | ···················· | ···················· |
| F | 5555 | hCoV-19/SaudiArabia/KAUST-MADINAH274/2020\|EPI_ISL_ | ···················· | ···················· | ···················· | ···················· | ···················· | ···················· |
| F | 5556 | hCoV-19/SaudiArabia/KAUST-MADINAH276/2020\|EPI_ISL_ | ···················· | ···················· | ···················· | ···················· | ···················· | ···················· |
| F | 5557 | hCoV-19/SaudiArabia/KAUST-MADINAH280/2020\|EPI_ISL_ | ···················· | ···················· | ···················· | ···················· | ···················· | ···················· |
| F | 5558 | hCoV-19/Saudi Arabia/KAUST-MADINAH281/2020\|EPI_ISL | ···················· | ···················· | ···················· | ···················· | ···················· | ···················· |
| F | 5559 | hCoV-19/Saudi Arabia/KAUST-MADINAH282/2020\|EPI_ISL | ···················· | ···················· | ···················· | ···················· | ···················· | ···················· |
| F | 5560 | hCoV-19/SaudiArabia/KAUST-MADINAH333/2020\|EPI_ISL_ | ···················· | ···················· | ···················· | ···················· | ···················· | ···················· |
| F | 5561 | hCoV-19/SaudiArabia/KAUST-MADINAH335/2020\|EPI_ISL_ | ···················· | ···················· | ···················· | ···················· | ···················· | ···················· |
| F | 5562 | hCoV-19/SaudiArabia/KAUST-MADINAH342/2020\|EPI_ISL_ | ···················· | ···················· | ···················· | ···················· | ···················· | ···················· |
| F | 5563 | hCoV-19/Saudi Arabia/KAUST-MADINAH344/2020\|EPI_ISL | ···················· | ···················· | ···················· | ···················· | ···················· | ···················· |
| F | 5564 | hCoV-19/SaudiArabia/KAUST-MADINAH346/2020\|EPI_ISL_ | ···················· | ···················· | ···················· | ···················· | ···················· | ···················· |
| F | 5565 | hCoV-19/SaudiArabia/KAUST-MADINAH347/2020\|EPI_ISL_ | ···················· | ···················· | ···················· | ···················· | ···················· | ···················· |
| F | 5566 | hCoV-19/SaudiArabia/KAUST-MADINAH354/2020\|EPI_ISL_ | ···················· | ···················· | ···················· | ···················· | ···················· | ···················· |
| F | 5567 | hCoV-19/SaudiArabia/KAUST-MADINAH355/2020\|EPI_ISL_ | ···················· | ···················· | ···················· | ···················· | ···················· | ···················· |
| F | 5568 | hCoV-19/SaudiArabia/KAUST-MADINAH357/2020\|EPI_ISL_ | ···················· | ···················· | ···················· | ···················· | ···················· | ···················· |
| F | 5569 | hCoV-19/SaudiArabia/KAUST-MADINAH360/2020\|EPI_ISL_ | ···················· | ···················· | ···················· | ···················· | ···················· | ···················· |
| F | 5570 | hCoV-19/SaudiArabia/KAUST-MADINAH361/2020\|EPI_ISL_ | ···················· | ···················· | ···················· | ···················· | ···················· | ···················· |
| F | 5571 | hCoV-19/SaudiArabia/KAUST-MADINAH362/2020\|EPI_ISL_ | ···················· | ·A·················· | ···················· | ···················· | ···················· | ···A················ |
| F | 5572 | hCoV-19/SaudiArabia/KAUST-MADINAH364/2020\|EPI_ISL_ | ···················· | ···················· | ···················· | ···················· | ···················· | ···················· |
| F | 5573 | hCoV-19/SaudiArabia/KAUST-MADINAH368/2020\|EPI_ISL_ | ···················· | ···················· | ···················· | ···················· | ···················· | ···················· |
| F | 5574 | hCoV-19/SaudiArabia/KAUST-MADINAH378/2020\|EPI_ISL_ | ···················· | ·A·················· | ···················· | ···················· | ···················· | ···A················ |
| F | 5575 | hCoV-19/SaudiArabia/KAUST-MADINAH380/2020\|EPI_ISL_ | ···················· | ···················· | ···················· | ···················· | ···················· | ···················· |
| F | 5576 | hCoV-19/SaudiArabia/KAUST-MADINAH384/2020\|EPI_ISL_ | ···················· | ···················· | ···················· | ···················· | ···················· | ···················· |
| F | 5577 | hCoV-19/SaudiArabia/KAUST-MADINAH387/2020\|EPI_ISL_ | ···················· | ···················· | ···················· | ···················· | ···················· | ···A················ |
| F | 5578 | hCoV-19/SaudiArabia/KAUST-MADINAH391/2020\|EPI_ISL_ | ···················· | ···················· | ···················· | ···················· | ···················· | ···················· |
| F | 5579 | hCoV-19/SaudiArabia/KAUST-MADINAH393/2020\|EPI_ISL_ | ···················· | ···················· | ···················· | ···················· | ···················· | ···················· |
| F | 5580 | hCoV-19/SaudiArabia/KAUST-MADINAH396/2020\|EPI_ISL_ | ···················· | ·A·················· | ···················· | ···················· | ···················· | ···A················ |
| F | 5581 | hCoV-19/SaudiArabia/KAUST-MADINAH397/2020\|EPI_ISL_ | ···················· | ···················· | ···················· | ···················· | ···················· | ···················· |
| F | 5582 | hCoV-19/SaudiArabia/KAUST-MADINAH398/2020\|EPI_ISL_ | ···················· | ·A·················· | ···················· | ···················· | ···················· | ···A················ |
| F | 5583 | hCoV-19/SaudiArabia/KAUST-MADINAH404/2020\|EPI_ISL_ | ···················· | ···················· | ···················· | ···················· | ···················· | ···················· |
| F | 5584 | hCoV-19/SaudiArabia/KAUST-MADINAH406/2020\|EPI_ISL_ | ···················· | ···················· | ···················· | ···················· | ···················· | ···················· |
| F | 5585 | hCoV-19/SaudiArabia/KAUST-MADINAH435/2020\|EPI_ISL_ | ···················· | ···················· | ···················· | ···················· | ···················· | ···················· |
| F | 5586 | hCoV-19/SaudiArabia/KAUST-MADINAH437/2020\|EPI_ISL_ | ···················· | ···················· | ···················· | ···················· | ···················· | ···················· |
| F | 5587 | hCoV-19/SaudiArabia/KAUST-MADINAH444/2020\|EPI_ISL_ | ···················· | ···················· | ···················· | ···················· | ···················· | ···················· |
| F | 5588 | hCoV-19/SaudiArabia/KAUST-MADINAH446/2020\|EPI_ISL_ | ···················· | ···················· | ···················· | ···················· | ···················· | ···················· |
| F | 5589 | hCoV-19/USA/CA-CZB-2462/2020\|EPI_ISL_513867\|2020-0 | ···················· | ···················· | ···················· | ···················· | ···················· | ···················· |
| F | 5590 | hCoV-19/England/CAMB-1B5AB0/2020\|EPI_ISL_514450\|20 | ···················· | ···················· | ···················· | ···················· | ···················· | ···················· |
| F | 5591 | hCoV-19/England/CAMB-1B5ACF/2020\|EPI_ISL_514451\|20 | ···················· | ···················· | ···················· | ···················· | ···················· | ···················· |
| F | 5592 | hCoV-19/England/NOTT-112EB5/2020\|EPI_ISL_514452\|20 | ···················· | ···················· | ···················· | ···················· | ···················· | ···················· |

| | | | | | | | | |
|---|---|---|---|---|---|---|---|---|
| F | 5593 | hCoV-19/England/NOTT-112ED3/2020\|EPI_ISL_514453\|20 | ···················· | ···················· | ···················· | ···················· | ···················· | ···················· |
| F | 5594 | hCoV-19/England/NOTT-112EF1/2020\|EPI_ISL_514454\|20 | ···················· | ···················· | ···················· | ···················· | ···················· | ···················· |
| F | 5595 | hCoV-19/England/PORT-2D16AA/2020\|EPI_ISL_514457\|20 | ···················· | ···················· | ···················· | ···················· | ···················· | ···················· |
| F | 5596 | hCoV-19/England/PORT-2D16D7/2020\|EPI_ISL_514458\|20 | ···················· | ···················· | ···················· | ···················· | ···················· | ···················· |
| F | 5597 | hCoV-19/England/PORT-2D16E6/2020\|EPI_ISL_514459\|20 | ···················· | ···················· | ···················· | ···················· | ···················· | ···················· |
| F | 5598 | hCoV-19/England/PORT-2D16F5/2020\|EPI_ISL_514460\|20 | ···················· | ···················· | ···················· | ···················· | ···················· | ···················· |
| F | 5599 | hCoV-19/England/PORT-2D1701/2020\|EPI_ISL_514461\|20 | ···················· | ···················· | ···················· | ···················· | ···················· | ···················· |
| F | 5600 | hCoV-19/England/PORT-2D1710/2020\|EPI_ISL_514462\|20 | ···················· | ···················· | ···················· | ···················· | ···················· | ···················· |
| F | 5601 | hCoV-19/England/PORT-2D172F/2020\|EPI_ISL_514463\|20 | ···················· | ···················· | ···················· | ···················· | ···················· | ···················· |
| F | 5602 | hCoV-19/England/PORT-2D175C/2020\|EPI_ISL_514465\|20 | ···················· | ···················· | ···················· | ···················· | ···················· | ···················· |
| F | 5603 | hCoV-19/England/PORT-2D176B/2020\|EPI_ISL_514466\|20 | ···················· | ···················· | ···················· | ···················· | ···················· | ···················· |
| F | 5604 | hCoV-19/England/PORT-2D1789/2020\|EPI_ISL_514467\|20 | ···················· | ···················· | ···················· | ···················· | ···················· | ···················· |
| F | 5605 | hCoV-19/England/PORT-2D1798/2020\|EPI_ISL_514468\|20 | ···················· | ···················· | ···················· | ···················· | ···················· | ···················· |
| F | 5606 | hCoV-19/England/PORT-2D17A7/2020\|EPI_ISL_514469\|20 | ···················· | ···················· | ···················· | ···················· | ···················· | ···················· |
| F | 5607 | hCoV-19/England/PORT-2D1947/2020\|EPI_ISL_514472\|20 | ···················· | ···················· | ···················· | ···················· | ···················· | ···················· |
| F | 5608 | hCoV-19/England/PORT-2D1956/2020\|EPI_ISL_514473\|20 | ···················· | ···················· | ···················· | ···················· | ···················· | ···················· |
| F | 5609 | hCoV-19/England/PORT-2D1965/2020\|EPI_ISL_514474\|20 | ···················· | ···················· | ···················· | ···················· | ···················· | ···················· |
| F | 5610 | hCoV-19/England/PORT-2D1974/2020\|EPI_ISL_514475\|20 | ···················· | ···················· | ···················· | ···················· | ···················· | ···················· |
| F | 5611 | hCoV-19/England/PORT-2D1992/2020\|EPI_ISL_514476\|20 | ···················· | ···················· | ···················· | ···················· | ···················· | ···················· |
| F | 5612 | hCoV-19/England/PORT-2D19B0/2020\|EPI_ISL_514477\|20 | ···················· | ···················· | ···················· | ···················· | ···················· | ···················· |
| F | 5613 | hCoV-19/England/PORT-2D19CF/2020\|EPI_ISL_514478\|20 | ···················· | ···················· | ···················· | ···················· | ···················· | ···················· |
| F | 5614 | hCoV-19/England/PORT-2D19ED/2020\|EPI_ISL_514479\|20 | ···················· | ···················· | ···················· | ···················· | ···················· | ···················· |
| F | 5615 | hCoV-19/England/PORT-2D19FC/2020\|EPI_ISL_514480\|20 | ···················· | ···················· | ···················· | ···················· | ···················· | ···················· |
| F | 5616 | hCoV-19/England/PORT-2D1A17/2020\|EPI_ISL_514482\|20 | ···················· | ···················· | ···················· | ···················· | ···················· | ···················· |
| F | 5617 | hCoV-19/England/PORT-2D2014/2020\|EPI_ISL_514485\|20 | ···················· | ···················· | ···················· | ···················· | ···················· | ···················· |
| F | 5618 | hCoV-19/England/PORT-2D20E7/2020\|EPI_ISL_514486\|20 | ···················· | ····T··············· | ···················· | ···················· | ···················· | ···················· |
| F | 5619 | hCoV-19/South Africa/KRISP-K002418/2020\|EPI_ISL_51 | ···················· | ···················· | ···················· | ···················· | ···················· | ···················· |
| F | 5620 | hCoV-19/South Africa/KRISP-K002419/2020\|EPI_ISL_51 | ···················· | ···················· | ···················· | ···················· | ···················· | ···················· |
| F | 5621 | hCoV-19/South Africa/KRISP-K002420/2020\|EPI_ISL_51 | ···················· | ···················· | ···················· | ···················· | ···················· | ···················· |
| F | 5622 | hCoV-19/South Africa/KRISP-K002421/2020\|EPI_ISL_51 | ···················· | ···················· | ···················· | ···················· | ···················· | ···················· |
| F | 5623 | hCoV-19/South Africa/KRISP-K002423/2020\|EPI_ISL_51 | ···················· | ···················· | ···················· | ···················· | ···················· | ···················· |
| F | 5624 | hCoV-19/South Africa/KRISP-K002426/2020\|EPI_ISL_51 | ···················· | ···················· | ···················· | ···················· | ···················· | ···················· |
| F | 5625 | hCoV-19/South Africa/KRISP-K002428/2020\|EPI_ISL_51 | ···················· | ···················· | ···················· | ···················· | ···················· | ···················· |
| F | 5626 | hCoV-19/South Africa/KRISP-K002429/2020\|EPI_ISL_51 | ···················· | ···················· | ···················· | ···················· | ···················· | ···················· |
| F | 5627 | hCoV-19/South Africa/KRISP-K002430/2020\|EPI_ISL_51 | ···················· | ···················· | ···················· | ···················· | ···················· | ···················· |
| F | 5628 | hCoV-19/South Africa/KRISP-K002431/2020\|EPI_ISL_51 | ···················· | ···················· | ···················· | ···················· | ···················· | ···················· |
| F | 5629 | hCoV-19/South Africa/KRISP-K002437/2020\|EPI_ISL_51 | ···················· | ···················· | ···················· | ···················· | ···················· | ···················· |
| F | 5630 | hCoV-19/South Africa/KRISP-K002440/2020\|EPI_ISL_51 | ···················· | ···················· | ···················· | ···················· | ···················· | ···················· |
| F | 5631 | hCoV-19/South Africa/KRISP-K002441/2020\|EPI_ISL_51 | ···················· | ···················· | ···················· | ···················· | ···················· | ···················· |
| F | 5633 | hCoV-19/USA/CT-UW-3129/2020\|EPI_ISL_500500\|2020-03 | ···················· | ···················· | ···················· | ···················· | ···················· | ···················· |
| F | 5634 | hCoV-19/USA/CT-UW-3422/2020\|EPI_ISL_500501\|2020-03 | ···················· | ···················· | ···················· | ···················· | ···················· | ···················· |
| F | 5635 | hCoV-19/USA/NY-UW-MAYO21/2020\|EPI_ISL_500503\|2020- | ···················· | ···················· | ···················· | ···················· | ···················· | ···················· |
| F | 5636 | hCoV-19/USA/NY-UW-MAYO23/2020\|EPI_ISL_500504\|2020- | ···················· | ···················· | ···················· | ···················· | ···················· | ···················· |
| F | 5637 | hCoV-19/USA/NY-UW-MAYO26/2020\|EPI_ISL_500505\|2020- | ···················· | ···················· | ···················· | ···················· | ···················· | ···················· |
| F | 5638 | hCoV-19/USA/NY-UW-MAYO27/2020\|EPI_ISL_500506\|2020- | ···················· | ···················· | ···················· | ···················· | ···················· | ···················· |
| F | 5639 | hCoV-19/USA/NY-UW-MAYO28/2020\|EPI_ISL_500507\|2020- | ···················· | ···················· | ···················· | ···················· | ···················· | ···················· |
| F | 5640 | hCoV-19/USA/CO-UW-MAYO33/2020\|EPI_ISL_500508\|2020- | ···················· | ···················· | ···················· | ···················· | ···················· | ···················· |
| F | 5641 | hCoV-19/USA/CO-UW-MAYO37/2020\|EPI_ISL_500509\|2020- | ···················· | ···················· | ···················· | ···················· | ···················· | ···················· |
| F | 5642 | hCoV-19/USA/CO-UW-MAYO39/2020\|EPI_ISL_500510\|2020- | ···················· | ···················· | ···················· | ···················· | ···················· | ···················· |
| F | 5643 | hCoV-19/USA/MA-UW-MAYO41/2020\|EPI_ISL_500511\|2020- | ···················· | ···················· | ···················· | ···················· | ···················· | ···················· |
| F | 5644 | hCoV-19/USA/WA-UW-MAYO57/2020\|EPI_ISL_500512\|2020- | ···················· | ···················· | ···················· | ···················· | ···················· | ···················· |
| F | 5645 | hCoV-19/USA/GA-UW-MAYO61/2020\|EPI_ISL_500513\|2020- | ···················· | ···················· | ···················· | ···················· | ···················· | ···················· |
| F | 5646 | hCoV-19/USA/GA-UW-MAYO62/2020\|EPI_ISL_500514\|2020- | ···················· | ···················· | ···················· | ···················· | ···················· | ···················· |
| F | 5647 | hCoV-19/USA/GA-UW-MAYO64/2020\|EPI_ISL_500515\|2020- | ···················· | ···················· | ···················· | ···················· | ···················· | ···················· |
| F | 5648 | hCoV-19/USA/MN-UW-MAYO72/2020\|EPI_ISL_500516\|2020- | ···················· | ···················· | ···················· | ···················· | ···················· | ···················· |
| F | 5649 | hCoV-19/USA/SD-UW-MAYO73/2020\|EPI_ISL_500517\|2020- | ···················· | ···················· | ···················· | ···················· | ···················· | ···················· |
| F | 5650 | hCoV-19/USA/MN-UW-MAYO79/2020\|EPI_ISL_500518\|2020- | ···················· | ···················· | ···················· | ···················· | ···················· | ···················· |
| F | 5651 | hCoV-19/USA/WI-UW-MAYO8/2020\|EPI_ISL_500519\|2020-0 | ···················· | ···················· | ···················· | ···················· | ···················· | ···················· |
| F | 5652 | hCoV-19/USA/LA-UW-MAYO86/2020\|EPI_ISL_500520\|2020- | ···················· | ···················· | ···················· | ···················· | ···················· | ···················· |
| F | 5653 | hCoV-19/USA/LA-UW-MAYO87/2020\|EPI_ISL_500521\|2020- | ···················· | ···················· | ···················· | ···················· | ···················· | ···················· |
| F | 5654 | hCoV-19/USA/AL-UW-MAYO12/2020\|EPI_ISL_500522\|2020- | ···················· | ···················· | ···················· | ···················· | ···················· | ···················· |
| F | 5655 | hCoV-19/USA/KY-UW-MAYO14/2020\|EPI_ISL_500523\|2020- | ···················· | ···················· | ···················· | ···················· | ···················· | ···················· |
| F | 5656 | hCoV-19/USA/KY-UW-MAYO16/2020\|EPI_ISL_500524\|2020- | ···················· | ···················· | ···················· | ···················· | ···················· | ···················· |
| F | 5657 | hCoV-19/USA/NY-UW-MAYO24/2020\|EPI_ISL_500525\|2020- | ···················· | ···················· | ···················· | ···················· | ···················· | ···················· |
| F | 5658 | hCoV-19/USA/CO-UW-MAYO35/2020\|EPI_ISL_500526\|2020- | ···················· | ···················· | ···················· | ···················· | ···················· | ···················· |
| F | 5659 | hCoV-19/USA/CO-UW-MAYO38/2020\|EPI_ISL_500527\|2020- | ···················· | ···················· | ···················· | ···················· | ···················· | ···················· |
| F | 5660 | hCoV-19/USA/CO-UW-MAYO40/2020\|EPI_ISL_500528\|2020- | ···················· | ···················· | ···················· | ···················· | ···················· | ···················· |
| F | 5661 | hCoV-19/USA/MA-UW-MAYO45/2020\|EPI_ISL_500529\|2020- | ···················· | ···················· | ···················· | ···················· | ···················· | ···················· |
| F | 5662 | hCoV-19/USA/VT-UW-MAYO48/2020\|EPI_ISL_500530\|2020- | ···················· | ···················· | ···················· | ···················· | ···················· | ···················· |
| F | 5663 | hCoV-19/USA/VT-UW-MAYO50/2020\|EPI_ISL_500531\|2020- | ···················· | ···················· | ···················· | ···················· | ···················· | ···················· |
| F | 5664 | hCoV-19/USA/CA-UW-MAYO53/2020\|EPI_ISL_500533\|2020- | ···················· | ···················· | ···················· | ···················· | ···················· | ···················· |
| F | 5665 | hCoV-19/USA/CA-UW-MAYO55/2020\|EPI_ISL_500534\|2020- | ···················· | ···················· | ···················· | ···················· | ···················· | ···················· |
| F | 5666 | hCoV-19/USA/IL-UW-MAYO6/2020\|EPI_ISL_500535\|2020-0 | ···················· | ···················· | ···················· | ···················· | ···················· | ···················· |
| F | 5667 | hCoV-19/USA/MD-UW-MAYO66/2020\|EPI_ISL_500536\|2020- | ···················· | ···················· | ···················· | ···················· | ···················· | ···················· |
| F | 5668 | hCoV-19/USA/MN-UW-MAYO74/2020\|EPI_ISL_500537\|2020- | ···················· | ···················· | ···················· | ···················· | ···················· | ···················· |
| F | 5669 | hCoV-19/USA/LA-UW-MAYO88/2020\|EPI_ISL_500538\|2020- | ···················· | ···················· | ···················· | ···················· | ···················· | ···················· |
| F | 5670 | hCoV-19/Singapore/733/2020\|EPI_ISL_500539\|2020-06- | ···················· | ···················· | ···················· | ···················· | ···················· | ···················· |
| F | 5671 | hCoV-19/Singapore/734/2020\|EPI_ISL_500540\|2020-06- | ···················· | ···················· | ···················· | ···················· | ···················· | ···················· |
| F | 5672 | hCoV-19/Singapore/735/2020\|EPI_ISL_500541\|2020-04- | ···················· | ··········T········· | ···················· | ···················· | ···················· | ···················· |
| F | 5673 | hCoV-19/Singapore/721/2020\|EPI_ISL_500542\|2020-06- | ···················· | ···················· | ···················· | ···················· | ···················· | ···················· |

| | | | | | | | | |
|---|---|---|---|---|---|---|---|---|
| F | 5674 | hCoV-19/Singapore/722/2020\|EPI_ISL_500543\|2020-06- | ···················· | ···················· | ···················· | ···················· | ···················· | ···················· |
| F | 5675 | hCoV-19/Singapore/723/2020\|EPI_ISL_500544\|2020-06- | ···················· | ···················· | ···················· | ···················· | ···················· | ···················· |
| F | 5676 | hCoV-19/Singapore/724/2020\|EPI_ISL_500545\|2020-06- | ···················· | ···················· | ···················· | ···················· | ···················· | ···················· |
| F | 5677 | hCoV-19/Singapore/725/2020\|EPI_ISL_500546\|2020-06- | ···················· | ···················· | ···················· | ···················· | ···················· | ···················· |
| F | 5678 | hCoV-19/Singapore/726/2020\|EPI_ISL_500547\|2020-06- | ···················· | ···················· | ···················· | ···················· | ···················· | ···················· |
| F | 5679 | hCoV-19/Singapore/727/2020\|EPI_ISL_500548\|2020-06- | ···················· | ···················· | ···················· | ···················· | ···················· | ···················· |
| F | 5680 | hCoV-19/Singapore/729/2020\|EPI_ISL_500550\|2020-06- | ···················· | ···················· | ···················· | ···················· | ···················· | ···················· |
| F | 5681 | hCoV-19/Singapore/730/2020\|EPI_ISL_500551\|2020-06- | ···················· | ···················· | ···················· | ···················· | ···················· | ···················· |
| F | 5682 | hCoV-19/Singapore/731/2020\|EPI_ISL_500552\|2020-06- | ···················· | ···················· | ···················· | ···················· | ···················· | ···················· |
| F | 5683 | hCoV-19/Singapore/732/2020\|EPI_ISL_500553\|2020-06- | ···················· | ···················· | ···················· | ···················· | ···················· | ···················· |
| F | 5684 | hCoV-19/Singapore/744/2020\|EPI_ISL_500554\|2020-06- | ···················· | ···················· | ···················· | ···················· | ···················· | ···················· |
| F | 5685 | hCoV-19/Singapore/601nan/2020\|EPI_ISL_500555\|2020- | ···················· | ···················· | ···················· | ···················· | ···················· | ···················· |
| F | 5686 | hCoV-19/Singapore/742/2020\|EPI_ISL_500556\|2020-07- | ···················· | ···················· | ···················· | ···················· | ···················· | ···················· |
| F | 5687 | hCoV-19/Singapore/737/2020\|EPI_ISL_500557\|2020-07- | ···················· | ···················· | ···················· | ···················· | ···················· | ···················· |
| F | 5688 | hCoV-19/Singapore/738/2020\|EPI_ISL_500558\|2020-07- | ···················· | ···················· | ···················· | ···················· | ···················· | ···················· |
| F | 5689 | hCoV-19/Singapore/736/2020\|EPI_ISL_500559\|2020-07- | ···················· | ···················· | ···················· | ···················· | ···················· | ···················· |
| F | 5690 | hCoV-19/Singapore/746/2020\|EPI_ISL_500560\|2020-07- | ···················· | ···················· | ···················· | ···················· | ···················· | ···················· |
| F | 5691 | hCoV-19/Singapore/747/2020\|EPI_ISL_500561\|2020-07- | ···················· | ···················· | ···················· | ···················· | ···················· | ···················· |
| F | 5692 | hCoV-19/Singapore/748/2020\|EPI_ISL_500562\|2020-06- | ···················· | ···················· | ···················· | ···················· | ···················· | ···················· |
| F | 5693 | hCoV-19/Singapore/751/2020\|EPI_ISL_500563\|2020-06- | ···················· | ···················· | ···················· | ···················· | ···················· | ···················· |
| F | 5694 | hCoV-19/Singapore/749/2020\|EPI_ISL_500564\|2020-06- | ···················· | ···················· | ···················· | ···················· | ···················· | ···················· |
| F | 5695 | hCoV-19/Singapore/750/2020\|EPI_ISL_500565\|2020-06- | ···················· | ···················· | ···················· | ···················· | ···················· | ···················· |
| F | 5696 | hCoV-19/Singapore/752/2020\|EPI_ISL_500566\|2020-06- | ···················· | ···················· | ···················· | ···················· | ···················· | ···················· |
| F | 5697 | hCoV-19/Singapore/753/2020\|EPI_ISL_500567\|2020-06- | ···················· | ···················· | ···················· | ···················· | ···················· | ···················· |
| F | 5698 | hCoV-19/Singapore/756/2020\|EPI_ISL_500570\|2020-06- | ···················· | ···················· | ···················· | ···················· | ···················· | ···················· |
| F | 5699 | hCoV-19/Ireland/WW-NVRL-71IRL07442/2020\|EPI_ISL_50 | ···················· | ·A·················· | ···················· | ···················· | ···················· | ···A················ |
| F | 5700 | hCoV-19/Ireland/CO-NVRL-71IRL17107/2020\|EPI_ISL_50 | ···················· | ···················· | ···················· | ···················· | ···················· | ···················· |
| F | 5702 | hCoV-19/Ireland/KY-NVRL-71IRL24965/2020\|EPI_ISL_50 | ···················· | ·A·················· | ···················· | ···················· | ···················· | ···A················ |
| F | 5703 | hCoV-19/Australia/NSW2004/2020\|EPI_ISL_500596\|2020 | ···················· | ···················· | ···················· | ···················· | ···················· | ···················· |
| F | 5704 | hCoV-19/Australia/NSW2005/2020\|EPI_ISL_500597\|2020 | ···················· | ···················· | ···················· | ···················· | ···················· | ···················· |
| F | 5705 | hCoV-19/Australia/NSW2011/2020\|EPI_ISL_500598\|2020 | ···················· | ···················· | ···················· | ···················· | ···················· | ···················· |
| F | 5706 | hCoV-19/Australia/NSW2013/2020\|EPI_ISL_500599\|2020 | ···················· | ···················· | ···················· | ···················· | ···················· | ···················· |
| F | 5707 | hCoV-19/Latvia/075/2020\|EPI_ISL_501817\|2020-07-09 | ···················· | ···················· | ···················· | ···················· | ···················· | ···················· |
| F | 5708 | hCoV-19/Latvia/076/2020\|EPI_ISL_501823\|2020-07-16 | ···················· | ···················· | ···················· | ···················· | ···················· | ···················· |
| F | 5709 | hCoV-19/Latvia/077/2020\|EPI_ISL_501829\|2020-07-16 | ···················· | ···················· | ···················· | ···················· | ···················· | ···················· |
| F | 5710 | hCoV-19/Latvia/078/2020\|EPI_ISL_501833\|2020-07-17 | ···················· | ···················· | ···················· | ···················· | ···················· | ···················· |
| F | 5711 | hCoV-19/Latvia/079/2020\|EPI_ISL_501839\|2020-07-17 | ···················· | ···················· | ···················· | ···················· | ···················· | ···················· |
| F | 5712 | hCoV-19/Latvia/080/2020\|EPI_ISL_501849\|2020-07-18 | ···················· | ···················· | ···················· | ···················· | ···················· | ···················· |
| F | 5713 | hCoV-19/Latvia/081/2020\|EPI_ISL_501894\|2020-07-20 | ···················· | ···················· | ···················· | ···················· | ···················· | ···················· |
| F | 5714 | hCoV-19/Latvia/082/2020\|EPI_ISL_501895\|2020-07-14 | ···················· | ···················· | ···················· | ···················· | ···················· | ···················· |
| F | 5715 | hCoV-19/Latvia/083/2020\|EPI_ISL_501896\|2020-06-15 | ···················· | ···················· | ···················· | ···················· | ···················· | ···················· |
| F | 5716 | hCoV-19/Portugal/PT1239/2020\|EPI_ISL_511200\|2020-0 | ···················· | ···················· | ···················· | ···················· | ···················· | ···················· |
| F | 5717 | hCoV-19/Portugal/PT1240/2020\|EPI_ISL_511201\|2020-0 | ···················· | ···················· | ···················· | ···················· | ···················· | ···················· |
| F | 5718 | hCoV-19/Portugal/PT1241/2020\|EPI_ISL_511202\|2020-0 | ···················· | ···················· | ···················· | ···················· | ···················· | ···················· |
| F | 5719 | hCoV-19/Portugal/PT1242/2020\|EPI_ISL_511203\|2020-0 | ···················· | ···················· | ···················· | ···················· | ···················· | ···················· |
| F | 5720 | hCoV-19/Portugal/PT1243/2020\|EPI_ISL_511204\|2020-0 | ···················· | ···················· | ···················· | ···················· | ···················· | ···················· |
| F | 5721 | hCoV-19/Portugal/PT1246/2020\|EPI_ISL_511207\|2020-0 | ···················· | ···················· | ···················· | ···················· | ···················· | ···················· |
| F | 5722 | hCoV-19/Portugal/PT1247/2020\|EPI_ISL_511208\|2020-0 | ···················· | ···················· | ···················· | ···················· | ···················· | ···················· |
| F | 5723 | hCoV-19/Portugal/PT1250/2020\|EPI_ISL_511211\|2020-0 | ···················· | ···················· | ···················· | ···················· | ···················· | ···················· |
| F | 5724 | hCoV-19/Portugal/PT1251/2020\|EPI_ISL_511212\|2020-0 | ···················· | ···················· | ···················· | ···················· | ···················· | ···················· |
| F | 5725 | hCoV-19/Portugal/PT1252/2020\|EPI_ISL_511213\|2020-0 | ···················· | ···················· | ···················· | ···················· | ···················· | ···················· |
| F | 5727 | hCoV-19/Portugal/PT1254/2020\|EPI_ISL_511215\|2020-0 | ···················· | ···················· | ···················· | ···················· | ···················· | ···················· |
| F | 5728 | hCoV-19/Portugal/PT1277/2020\|EPI_ISL_511238\|2020-0 | ···················· | ···················· | ···················· | ···················· | ···················· | ···················· |
| F | 5729 | hCoV-19/Portugal/PT1278/2020\|EPI_ISL_511239\|2020-0 | ···················· | ···················· | ···················· | ···················· | ···················· | ···················· |
| F | 5730 | hCoV-19/Portugal/PT1279/2020\|EPI_ISL_511240\|2020-0 | ···················· | ···················· | ···················· | ···················· | ···················· | ···················· |
| F | 5731 | hCoV-19/Portugal/PT1280/2020\|EPI_ISL_511241\|2020-0 | ···················· | ···················· | ···················· | ···················· | ···················· | ···················· |
| F | 5732 | hCoV-19/Portugal/PT1281/2020\|EPI_ISL_511242\|2020-0 | ···················· | ···················· | ···················· | ···················· | ···················· | ···················· |
| F | 5733 | hCoV-19/Portugal/PT1282/2020\|EPI_ISL_511243\|2020-0 | ···················· | ···················· | ···················· | ···················· | ···················· | ···················· |
| F | 5735 | hCoV-19/Portugal/PT1284/2020\|EPI_ISL_511245\|2020-0 | ···················· | ···················· | ···················· | ···················· | ···················· | ···················· |
| F | 5736 | hCoV-19/Portugal/PT1285/2020\|EPI_ISL_511246\|2020-0 | ···················· | ···················· | ···················· | ···················· | ···················· | ···················· |
| F | 5738 | hCoV-19/Portugal/PT1287/2020\|EPI_ISL_511248\|2020-0 | ···················· | ···················· | ···················· | ···················· | ···················· | ···················· |
| F | 5739 | hCoV-19/Portugal/PT1288/2020\|EPI_ISL_511249\|2020-0 | ···················· | ···················· | ···················· | ···················· | ···················· | ···················· |
| F | 5740 | hCoV-19/Portugal/PT1289/2020\|EPI_ISL_511250\|2020-0 | ···················· | ···················· | ···················· | ···················· | ···················· | ···················· |
| F | 5741 | hCoV-19/Portugal/PT1290/2020\|EPI_ISL_511251\|2020-0 | ···················· | ···················· | ···················· | ···················· | ···················· | ···················· |
| F | 5742 | hCoV-19/Portugal/PT1291/2020\|EPI_ISL_511252\|2020-0 | ···················· | ···················· | ···················· | ···················· | ···················· | ···················· |
| F | 5743 | hCoV-19/Portugal/PT1292/2020\|EPI_ISL_511253\|2020-0 | ···················· | ···················· | ···················· | ···················· | ···················· | ···················· |
| F | 5744 | hCoV-19/Portugal/PT1293/2020\|EPI_ISL_511254\|2020-0 | ···················· | ···················· | ···················· | ···················· | ···················· | ···················· |
| F | 5745 | hCoV-19/Portugal/PT1294/2020\|EPI_ISL_511255\|2020-0 | ···················· | ···················· | ···················· | ···················· | ···················· | ···················· |
| F | 5747 | hCoV-19/Portugal/PT1300/2020\|EPI_ISL_511259\|2020-0 | ···················· | ···················· | ···················· | ···················· | ···················· | ···················· |
| F | 5748 | hCoV-19/Portugal/PT1302/2020\|EPI_ISL_511261\|2020-0 | ···················· | ···················· | ···················· | ···················· | ···················· | ···················· |
| F | 5749 | hCoV-19/Portugal/PT1305/2020\|EPI_ISL_511264\|2020-0 | ···················· | ···················· | ···················· | ···················· | ···················· | ···················· |
| F | 5750 | hCoV-19/Portugal/PT1308/2020\|EPI_ISL_511267\|2020-0 | ···················· | ···················· | ···················· | ···················· | ···················· | ···················· |
| F | 5751 | hCoV-19/Portugal/PT1309/2020\|EPI_ISL_511268\|2020-0 | ···················· | ···················· | ···················· | ···················· | ···················· | ···················· |
| F | 5752 | hCoV-19/Portugal/PT1310/2020\|EPI_ISL_511269\|2020-0 | ···················· | ···················· | ···················· | ···················· | ···················· | ···················· |
| F | 5753 | hCoV-19/Portugal/PT1312/2020\|EPI_ISL_511271\|2020-0 | ···················· | ···················· | ···················· | ···················· | ···················· | ···················· |
| F | 5754 | hCoV-19/Portugal/PT1313/2020\|EPI_ISL_511272\|2020-0 | ···················· | ···················· | ···················· | ···················· | ···················· | ···················· |
| F | 5755 | hCoV-19/Portugal/PT1315/2020\|EPI_ISL_511274\|2020-0 | ···················· | ···················· | ···················· | ···················· | ···················· | ···················· |
| F | 5756 | hCoV-19/Portugal/PT1316/2020\|EPI_ISL_511275\|2020-0 | ···················· | ···················· | ···················· | ···················· | ···················· | ···················· |
| F | 5757 | hCoV-19/Portugal/PT1317/2020\|EPI_ISL_511276\|2020-0 | ···················· | ···················· | ···················· | ···················· | ···················· | ···················· |
| F | 5758 | hCoV-19/Portugal/PT1318/2020\|EPI_ISL_511277\|2020-0 | ···················· | ···················· | ···················· | ···················· | ···················· | ···················· |

| | | | | | | | | |
|---|---|---|---|---|---|---|---|---|
| F | 5759 | hCoV-19/Portugal/PT1322/2020\|EPI_ISL_511281\|2020-0 | .................... | .................... | .................... | .................... | .................... | .................... |
| F | 5760 | hCoV-19/Portugal/PT1324/2020\|EPI_ISL_511283\|2020-0 | .................... | .................... | .................... | .................... | .................... | .................... |
| F | 5761 | hCoV-19/Portugal/PT1325/2020\|EPI_ISL_511284\|2020-0 | .................... | .................... | .................... | .................... | .................... | .................... |
| F | 5762 | hCoV-19/Portugal/PT1326/2020\|EPI_ISL_511285\|2020-0 | .................... | .................... | .................... | .................... | .................... | .................... |
| F | 5763 | hCoV-19/Portugal/PT1327/2020\|EPI_ISL_511286\|2020-0 | .................... | .................... | .................... | .................... | .................... | .................... |
| F | 5764 | hCoV-19/Portugal/PT1328/2020\|EPI_ISL_511287\|2020-0 | .................... | .................... | .................... | .................... | .................... | .................... |
| F | 5765 | hCoV-19/Portugal/PT1329/2020\|EPI_ISL_511288\|2020-0 | .................... | .................... | .................... | .................... | .................... | .................... |
| F | 5766 | hCoV-19/Portugal/PT1330/2020\|EPI_ISL_511289\|2020-0 | .................... | .................... | .................... | .................... | .................... | .................... |
| F | 5767 | hCoV-19/Portugal/PT1331/2020\|EPI_ISL_511290\|2020-0 | .................... | .................... | .................... | .................... | .................... | .................... |
| F | 5768 | hCoV-19/Portugal/PT1335/2020\|EPI_ISL_511294\|2020-0 | .................... | .................... | .................... | .................... | .................... | .................... |
| F | 5769 | hCoV-19/Portugal/PT1336/2020\|EPI_ISL_511295\|2020-0 | .................... | .................... | .................... | .................... | .................... | .................... |
| F | 5770 | hCoV-19/Portugal/PT1337/2020\|EPI_ISL_511296\|2020-0 | .................... | .................... | .................... | .................... | .................... | .................... |
| F | 5771 | hCoV-19/Portugal/PT1338/2020\|EPI_ISL_511297\|2020-0 | .................... | .................... | .................... | .................... | .................... | .................... |
| F | 5772 | hCoV-19/Portugal/PT1339/2020\|EPI_ISL_511298\|2020-0 | .................... | .................... | .................... | .................... | .................... | .................... |
| F | 5773 | hCoV-19/Portugal/PT1340/2020\|EPI_ISL_511299\|2020-0 | .................... | .................... | .................... | .................... | .................... | .................... |
| F | 5775 | hCoV-19/Wales/PHWC-1696F3/2020\|EPI_ISL_512501\|2020 | .................... | .................... | .................... | .................... | .................... | .................... |
| F | 5777 | hCoV-19/Wales/PHWC-16971E/2020\|EPI_ISL_512503\|2020 | .................... | .................... | .................... | .................... | .................... | .................... |
| F | 5778 | hCoV-19/Wales/PHWC-16972D/2020\|EPI_ISL_512504\|2020 | .................... | .................... | .................... | .................... | .................... | .................... |
| F | 5779 | hCoV-19/Wales/PHWC-16973C/2020\|EPI_ISL_512505\|2020 | .................... | .................... | .................... | .................... | .................... | .................... |
| F | 5780 | hCoV-19/Wales/PHWC-16974B/2020\|EPI_ISL_512506\|2020 | .................... | .................... | .................... | .................... | .................... | .................... |
| F | 5781 | hCoV-19/Wales/PHWC-16975A/2020\|EPI_ISL_512507\|2020 | .................... | .................... | .................... | .................... | .................... | .................... |
| F | 5782 | hCoV-19/Wales/PHWC-169769/2020\|EPI_ISL_512508\|2020 | .................... | .................... | .................... | .................... | .................... | .................... |
| F | 5783 | hCoV-19/Wales/PHWC-169778/2020\|EPI_ISL_512509\|2020 | .................... | .................... | .................... | .................... | .................... | .................... |
| F | 5784 | hCoV-19/Wales/PHWC-169796/2020\|EPI_ISL_512510\|2020 | .................... | .................... | .................... | .................... | .................... | .................... |
| F | 5785 | hCoV-19/Wales/PHWC-1697A5/2020\|EPI_ISL_512511\|2020 | .................... | .................... | .................... | .................... | .................... | .................... |
| F | 5786 | hCoV-19/Wales/PHWC-1697C3/2020\|EPI_ISL_512513\|2020 | .................... | .................... | .................... | .................... | .................... | .................... |
| F | 5787 | hCoV-19/Wales/PHWC-1697E1/2020\|EPI_ISL_512515\|2020 | .................... | .................... | .................... | .................... | .................... | .................... |
| F | 5788 | hCoV-19/Wales/PHWC-16980C/2020\|EPI_ISL_512517\|2020 | .................... | .................... | .................... | .................... | .................... | .................... |
| F | 5789 | hCoV-19/Wales/PHWC-16981B/2020\|EPI_ISL_512518\|2020 | .................... | .................... | .................... | .................... | .................... | .................... |
| F | 5790 | hCoV-19/Wales/PHWC-16982A/2020\|EPI_ISL_512519\|2020 | .................... | .................... | .................... | .................... | .................... | .................... |
| F | 5791 | hCoV-19/Wales/PHWC-169839/2020\|EPI_ISL_512520\|2020 | .................... | .................... | .................... | .................... | .................... | .................... |
| F | 5792 | hCoV-19/Wales/PHWC-169848/2020\|EPI_ISL_512521\|2020 | .................... | .................... | .................... | .................... | .................... | .................... |
| F | 5793 | hCoV-19/Wales/PHWC-169857/2020\|EPI_ISL_512522\|2020 | .................... | .................... | .................... | .................... | .................... | .................... |
| F | 5794 | hCoV-19/Wales/PHWC-169866/2020\|EPI_ISL_512523\|2020 | .................... | .................... | .................... | .................... | .................... | .................... |
| F | 5795 | hCoV-19/Wales/PHWC-169875/2020\|EPI_ISL_512524\|2020 | .................... | .................... | .................... | .................... | .................... | .................... |
| F | 5796 | hCoV-19/Wales/PHWC-1698A2/2020\|EPI_ISL_512527\|2020 | .................... | .................... | .................... | .................... | .................... | .................... |
| F | 5797 | hCoV-19/Wales/PHWC-1698B1/2020\|EPI_ISL_512528\|2020 | .................... | .................... | .................... | .................... | .................... | .................... |
| F | 5798 | hCoV-19/Wales/PHWC-1698EE/2020\|EPI_ISL_512531\|2020 | .................... | .................... | .................... | .................... | .................... | .................... |
| F | 5799 | hCoV-19/Wales/PHWC-1698FD/2020\|EPI_ISL_512532\|2020 | .................... | .................... | .................... | .................... | .................... | .................... |
| F | 5800 | hCoV-19/Wales/PHWC-169909/2020\|EPI_ISL_512533\|2020 | .................... | .................... | .................... | .................... | .................... | .................... |
| F | 5801 | hCoV-19/Wales/PHWC-169927/2020\|EPI_ISL_512535\|2020 | .................... | .................... | .................... | .................... | .................... | .................... |
| F | 5802 | hCoV-19/Wales/PHWC-169954/2020\|EPI_ISL_512538\|2020 | .................... | .................... | .................... | .................... | .................... | .................... |
| F | 5803 | hCoV-19/Wales/PHWC-169972/2020\|EPI_ISL_512540\|2020 | .................... | .................... | .................... | .................... | .................... | .................... |
| F | 5804 | hCoV-19/Wales/PHWC-169981/2020\|EPI_ISL_512541\|2020 | .................... | .................... | .................... | .................... | .................... | .................... |
| F | 5805 | hCoV-19/Wales/PHWC-169990/2020\|EPI_ISL_512542\|2020 | .................... | .................... | .................... | .................... | .................... | .................... |
| F | 5806 | hCoV-19/Wales/PHWC-1699AF/2020\|EPI_ISL_512543\|2020 | .................... | .................... | .................... | .................... | .................... | .................... |
| F | 5807 | hCoV-19/Wales/PHWC-1699BE/2020\|EPI_ISL_512544\|2020 | .................... | .................... | .................... | .................... | .................... | .................... |
| F | 5808 | hCoV-19/Wales/PHWC-1699CD/2020\|EPI_ISL_512545\|2020 | .................... | .................... | .................... | .................... | .................... | .................... |
| F | 5809 | hCoV-19/USA/FL-BPHL-0752/2020\|EPI_ISL_512558\|2020- | .................... | .................... | .................... | .................... | .................... | .................... |
| F | 5811 | hCoV-19/USA/FL-BPHL-0765/2020\|EPI_ISL_512570\|2020- | .................... | .................... | .................... | .................... | .................... | .................... |
| F | 5812 | hCoV-19/USA/FL-BPHL-0766/2020\|EPI_ISL_512571\|2020- | .................... | .................... | .................... | .................... | .................... | .................... |
| F | 5813 | hCoV-19/USA/FL-BPHL-0767/2020\|EPI_ISL_512572\|2020- | .................... | .................... | .................... | .................... | .................... | .................... |
| F | 5814 | hCoV-19/USA/FL-BPHL-0768/2020\|EPI_ISL_512573\|2020- | .................... | .................... | .................... | .................... | .................... | .................... |
| F | 5815 | hCoV-19/USA/FL-BPHL-0770/2020\|EPI_ISL_512575\|2020- | .................... | .................... | .................... | .................... | .................... | .................... |
| F | 5816 | hCoV-19/USA/FL-BPHL-0774/2020\|EPI_ISL_512578\|2020- | .................... | .................... | .................... | .................... | .................... | .................... |
| F | 5818 | hCoV-19/Ukraine/203100317/2020\|EPI_ISL_512597\|2020 | .................... | .................... | .................... | .................... | .................... | .................... |
| F | 5819 | hCoV-19/Ukraine/203100318/2020\|EPI_ISL_512598\|2020 | .................... | .................... | .................... | .................... | .................... | .................... |
| F | 5820 | hCoV-19/Ukraine/203100319/2020\|EPI_ISL_512599\|2020 | .................... | .................... | .................... | .................... | .................... | .................... |
| F | 5821 | hCoV-19/USA/CA-CZB-2353/2020\|EPI_ISL_513800\|2020-0 | .................... | .................... | .................... | .................... | .................... | .................... |
| F | 5822 | hCoV-19/USA/CA-CZB-2354/2020\|EPI_ISL_513801\|2020-0 | .................... | .................... | .................... | .................... | .................... | .................... |
| F | 5823 | hCoV-19/USA/CA-CZB-2355/2020\|EPI_ISL_513802\|2020-0 | .................... | .................... | .................... | .................... | .................... | .................... |
| F | 5824 | hCoV-19/USA/CA-CZB-2356/2020\|EPI_ISL_513803\|2020-0 | .................... | .................... | .................... | .................... | .................... | .................... |
| F | 5825 | hCoV-19/USA/CA-CZB-2357/2020\|EPI_ISL_513804\|2020-0 | .................... | .................... | .................... | .................... | .................... | .................... |
| F | 5826 | hCoV-19/USA/CA-CZB-2358/2020\|EPI_ISL_513805\|2020-0 | .................... | .................... | .................... | .................... | .................... | .................... |
| F | 5827 | hCoV-19/USA/CA-CZB-2360/2020\|EPI_ISL_513806\|2020-0 | .................... | .................... | .................... | .................... | .................... | .................... |
| F | 5828 | hCoV-19/USA/CA-CZB-2374/2020\|EPI_ISL_513814\|2020-0 | .................... | .................... | .................... | .................... | .................... | .................... |
| F | 5829 | hCoV-19/USA/CA-CZB-2375/2020\|EPI_ISL_513815\|2020-0 | .................... | .................... | .................... | .................... | .................... | .................... |
| F | 5830 | hCoV-19/USA/CA-CZB-2376/2020\|EPI_ISL_513816\|2020-0 | .................... | .................... | .................... | .................... | .................... | .................... |
| F | 5831 | hCoV-19/USA/CA-CZB-2377/2020\|EPI_ISL_513817\|2020-0 | .................... | .................... | .................... | .................... | .................... | .................... |
| F | 5832 | hCoV-19/USA/CA-CZB-2387/2020\|EPI_ISL_513820\|2020-0 | .................... | .................... | .................... | .................... | .................... | .................... |
| F | 5833 | hCoV-19/USA/CA-CZB-2391/2020\|EPI_ISL_513822\|2020-0 | .................... | .................... | .................... | .................... | .................... | .................... |
| F | 5834 | hCoV-19/USA/CA-CZB-2395/2020\|EPI_ISL_513824\|2020-0 | .................... | .................... | .................... | .................... | .................... | .................... |
| F | 5835 | hCoV-19/USA/CA-CZB-2397/2020\|EPI_ISL_513826\|2020-0 | .................... | .................... | .................... | .................... | .................... | .................... |
| F | 5836 | hCoV-19/USA/CA-CZB-2400/2020\|EPI_ISL_513828\|2020-0 | .................... | .................... | .................... | .................... | .................... | .................... |
| F | 5837 | hCoV-19/USA/CA-CZB-2401/2020\|EPI_ISL_513829\|2020-0 | .................... | .................... | .................... | .................... | .................... | .................... |
| F | 5838 | hCoV-19/USA/CA-CZB-2403/2020\|EPI_ISL_513830\|2020-0 | .................... | .................... | .................... | .................... | .................... | .................... |
| F | 5839 | hCoV-19/USA/CA-CZB-2408/2020\|EPI_ISL_513832\|2020-0 | .................... | .................... | .................... | .................... | .................... | .................... |
| F | 5840 | hCoV-19/USA/CA-CZB-2409/2020\|EPI_ISL_513833\|2020-0 | .................... | .................... | .................... | .................... | .................... | .................... |
| F | 5841 | hCoV-19/USA/CA-CZB-2413/2020\|EPI_ISL_513835\|2020-0 | .................... | .A.................. | .................... | .................... | .................... | .................... |
| F | 5842 | hCoV-19/USA/CA-CZB-2414/2020\|EPI_ISL_513836\|2020-0 | .................... | .A.................. | .................... | .................... | .................... | .................... |

| | | | | | | | |
|---|---|---|---|---|---|---|---|
| F | 5843 hCoV-19/USA/CA-CZB-2416/2020\|EPI_ISL_513837\|2020-0 | ···················· | ···················· | ···················· | ···················· | ···················· | ···················· |
| F | 5844 hCoV-19/USA/CA-CZB-2417/2020\|EPI_ISL_513838\|2020-0 | ···················· | ···················· | ···················· | ···················· | ···················· | ···················· |
| F | 5845 hCoV-19/USA/CA-CZB-2418/2020\|EPI_ISL_513839\|2020-0 | ···················· | ···················· | ···················· | ···················· | ···················· | ···················· |
| F | 5848 hCoV-19/USA/CA-CZB-2421/2020\|EPI_ISL_513842\|2020-0 | ···················· | ···················· | ···················· | ···················· | ···················· | ···················· |
| F | 5849 hCoV-19/USA/CA-CZB-2422/2020\|EPI_ISL_513843\|2020-0 | ···················· | ···················· | ···················· | ···················· | ···················· | ···················· |
| F | 5850 hCoV-19/USA/CA-CZB-2423/2020\|EPI_ISL_513844\|2020-0 | ···················· | ···················· | ···················· | ···················· | ···················· | ···················· |
| F | 5851 hCoV-19/USA/CA-CZB-2424/2020\|EPI_ISL_513845\|2020-0 | ···················· | ···················· | ···················· | ···················· | ···················· | ···················· |
| F | 5852 hCoV-19/USA/CA-CZB-2425/2020\|EPI_ISL_513846\|2020-0 | ···················· | ···················· | ···················· | ···················· | ···················· | ···················· |
| F | 5853 hCoV-19/USA/CA-CZB-2426/2020\|EPI_ISL_513847\|2020-0 | ···················· | ···················· | ···················· | ···················· | ···················· | ···················· |
| F | 5854 hCoV-19/USA/CA-CZB-2429/2020\|EPI_ISL_513848\|2020-0 | ···················· | ···················· | ···················· | ···················· | ···················· | ···················· |
| F | 5855 hCoV-19/USA/CA-CZB-2431/2020\|EPI_ISL_513850\|2020-0 | ···················· | ···················· | ···················· | ···················· | ···················· | ···················· |
| F | 5856 hCoV-19/USA/CA-CZB-2436/2020\|EPI_ISL_513852\|2020-0 | ···················· | ···················· | ···················· | ···················· | ···················· | ···················· |
| F | 5857 hCoV-19/USA/CA-CZB-2443/2020\|EPI_ISL_513853\|2020-0 | ···················· | ···················· | ···················· | ···················· | ···················· | ···················· |
| F | 5858 hCoV-19/USA/CA-CZB-2445/2020\|EPI_ISL_513854\|2020-0 | ···················· | ···················· | ···················· | ···················· | ···················· | ···················· |
| F | 5859 hCoV-19/USA/CA-CZB-2446/2020\|EPI_ISL_513855\|2020-0 | ···················· | ···················· | ···················· | ···················· | ···················· | ···················· |
| F | 5860 hCoV-19/USA/CA-CZB-2447/2020\|EPI_ISL_513856\|2020-0 | ···················· | ···················· | ···················· | ···················· | ···················· | ···················· |
| F | 5861 hCoV-19/USA/CA-CZB-2448/2020\|EPI_ISL_513857\|2020-0 | ···················· | ···················· | ···················· | ···················· | ···················· | ···················· |
| F | 5862 hCoV-19/USA/CA-CZB-2450/2020\|EPI_ISL_513858\|2020-0 | ···················· | ···················· | ···················· | ···················· | ···················· | ···················· |
| F | 5863 hCoV-19/USA/CA-CZB-2451/2020\|EPI_ISL_513859\|2020-0 | ···················· | ···················· | ···················· | ···················· | ···················· | ···················· |
| F | 5864 hCoV-19/USA/CA-CZB-2452/2020\|EPI_ISL_513860\|2020-0 | ···················· | ···················· | ···················· | ···················· | ···················· | ···················· |
| F | 5865 hCoV-19/USA/CA-CZB-2453/2020\|EPI_ISL_513861\|2020-0 | ···················· | ···················· | ···················· | ···················· | ···················· | ···················· |
| F | 5866 hCoV-19/USA/CA-CZB-2454/2020\|EPI_ISL_513862\|2020-0 | ···················· | ···················· | ···················· | ···················· | ···················· | ···················· |
| F | 5867 hCoV-19/USA/CA-CZB-2455/2020\|EPI_ISL_513863\|2020-0 | ···················· | ···················· | ···················· | ···················· | ···················· | ···················· |
| F | 5868 hCoV-19/USA/CA-CZB-2458/2020\|EPI_ISL_513864\|2020-0 | ···················· | ···················· | ···················· | ···················· | ···················· | ···················· |
| F | 5869 hCoV-19/USA/CA-CZB-2459/2020\|EPI_ISL_513865\|2020-0 | ···················· | ···················· | ···················· | ···················· | ···················· | ···················· |
| F | 5870 hCoV-19/USA/CA-CZB-2460/2020\|EPI_ISL_513866\|2020-0 | ···················· | ···················· | ···················· | ···················· | ···················· | ···················· |
| F | 5871 hCoV-19/USA/CA-CZB-2464/2020\|EPI_ISL_513869\|2020-0 | ···················· | ···················· | ···················· | ···················· | ···················· | ···················· |
| F | 5872 hCoV-19/USA/CA-CZB-2465/2020\|EPI_ISL_513870\|2020-0 | ···················· | ···················· | ···················· | ···················· | ···················· | ···················· |
| F | 5873 hCoV-19/USA/CA-CZB-2467/2020\|EPI_ISL_513872\|2020-0 | ···················· | ···················· | ···················· | ···················· | ···················· | ···················· |
| F | 5874 hCoV-19/USA/CA-CZB-2468/2020\|EPI_ISL_513873\|2020-0 | ···················· | ···················· | ···················· | ···················· | ···················· | ···················· |
| F | 5875 hCoV-19/USA/CA-CZB-2470/2020\|EPI_ISL_513874\|2020-0 | ···················· | ···················· | ···················· | ···················· | ···················· | ···················· |
| F | 5876 hCoV-19/USA/CA-CZB-2471/2020\|EPI_ISL_513875\|2020-0 | ···················· | ···················· | ···················· | ···················· | ···················· | ···················· |
| F | 5877 hCoV-19/USA/CA-CZB-2472/2020\|EPI_ISL_513876\|2020-0 | ···················· | ···················· | ···················· | ···················· | ···················· | ···················· |
| F | 5878 hCoV-19/USA/CA-CZB-2504/2020\|EPI_ISL_513889\|2020-0 | ···················· | ·A·················· | ···················· | ···················· | ···················· | ···················· |
| F | 5879 hCoV-19/Italy/MAR-UnivPM-32898/2020\|EPI_ISL_516079 | ···················· | ···················· | ···················· | ···················· | ···················· | ···················· |
| F | 5880 hCoV-19/Italy/MAR-UnivPM-34104/2020\|EPI_ISL_516080 | ···················· | ···················· | ···················· | ···················· | ···················· | ···················· |
| F | 5881 hCoV-19/Italy/MAR-UnivPM-34117/2020\|EPI_ISL_516081 | ···················· | ···················· | ···················· | ···················· | ···················· | ···················· |
| F | 5882 hCoV-19/Italy/MAR-UnivPM-43950/2020\|EPI_ISL_516082 | ···················· | ···················· | ···················· | ···················· | ···················· | ···················· |
| F | 5883 hCoV-19/Italy/MAR-UnivPM-71205/2020\|EPI_ISL_516083 | ···················· | ···················· | ···················· | ···················· | ···················· | ···················· |
| F | 5884 hCoV-19/Italy/MAR-UnivPM-73696/2020\|EPI_ISL_516084 | ···················· | ···················· | ···················· | ···················· | ···················· | ···················· |
| F | 5885 hCoV-19/Italy/MAR-UnivPM-73702/2020\|EPI_ISL_516085 | ···················· | ···················· | ···················· | ···················· | ···················· | ···················· |
| F | 5886 hCoV-19/Italy/MAR-UnivPM-78952/2020\|EPI_ISL_516086 | ···················· | ···················· | ···················· | ···················· | ···················· | ···················· |
| F | 5887 hCoV-19/Italy/MAR-UnivPM-78955/2020\|EPI_ISL_516087 | ···················· | ···················· | ···················· | ···················· | ···················· | ···················· |
| F | 5888 hCoV-19/Italy/MAR-UnivPM-78955-2/2020\|EPI_ISL_5160 | ···················· | ···················· | ···················· | ···················· | ···················· | ···················· |
| F | 5889 hCoV-19/Spain/RI-IBV-005966/2020\|EPI_ISL_500400\|20 | ···················· | ···················· | ···················· | ···················· | ···················· | ···················· |
| F | 5890 hCoV-19/Spain/RI-IBV-005967/2020\|EPI_ISL_500401\|20 | ···················· | ···················· | ···················· | ···················· | ···················· | ···················· |
| F | 5891 hCoV-19/Spain/RI-IBV-005968/2020\|EPI_ISL_500402\|20 | ···················· | ···················· | ···················· | ···················· | ···················· | ···················· |
| F | 5892 hCoV-19/Spain/RI-IBV-005973/2020\|EPI_ISL_500407\|20 | ···················· | ···················· | ···················· | ···················· | ···················· | ···················· |
| F | 5896 hCoV-19/Spain/RI-IBV-005977/2020\|EPI_ISL_500411\|20 | ···················· | ···················· | ···················· | ···················· | ···················· | ···················· |
| F | 5898 hCoV-19/Spain/RI-IBV-005979/2020\|EPI_ISL_500413\|20 | ···················· | ···················· | ···················· | ···················· | ···················· | ···················· |
| F | 5902 hCoV-19/Spain/RI-IBV-005987/2020\|EPI_ISL_500421\|20 | ···················· | ·A·················· | ···················· | ···················· | ···················· | ···················· |
| F | 5903 hCoV-19/Spain/RI-IBV-005990/2020\|EPI_ISL_500423\|20 | ···················· | ···················· | ···················· | ···················· | ···················· | ···················· |
| F | 5905 hCoV-19/Spain/RI-IBV-005995/2020\|EPI_ISL_500427\|20 | ···················· | ···················· | ···················· | ···················· | ··N················· | ···················· |
| F | 5906 hCoV-19/Spain/RI-IBV-005996/2020\|EPI_ISL_500428\|20 | ···················· | ···················· | ···················· | ···················· | ···················· | ···················· |
| F | 5907 hCoV-19/Spain/RI-IBV-005997/2020\|EPI_ISL_500429\|20 | ···················· | ···················· | ···················· | ···················· | ···················· | ···················· |
| F | 5908 hCoV-19/Spain/RI-IBV-005998/2020\|EPI_ISL_500430\|20 | ···················· | ···················· | ···················· | ···················· | ··NN················ | ···················· |
| F | 5909 hCoV-19/Spain/RI-IBV-005999/2020\|EPI_ISL_500431\|20 | ···················· | ···················· | ···················· | ···················· | ···················· | ···················· |
| F | 5910 hCoV-19/Spain/RI-IBV-006002/2020\|EPI_ISL_500433\|20 | ···················· | ···················· | ···················· | ···················· | ···················· | ···················· |
| F | 5913 hCoV-19/Singapore/754/2020\|EPI_ISL_500568\|2020-06- | ···················· | ···················· | ···················· | ···················· | ···················· | ···················· |
| F | 5914 hCoV-19/Singapore/755/2020\|EPI_ISL_500569\|2020-06- | ···················· | ···················· | ···················· | ···················· | ···················· | ···················· |
| F | 5915 hCoV-19/Singapore/757/2020\|EPI_ISL_500571\|2020-06- | ···················· | ···················· | ···················· | ···················· | ···················· | ···················· |
| F | 5916 hCoV-19/Singapore/745/2020\|EPI_ISL_500572\|2020-07- | ···················· | ···················· | ···················· | ···················· | ···················· | ···················· |
| F | 5917 hCoV-19/Ireland/SO-NVRL-70IRL90620/2020\|EPI_ISL_50 | ···················· | ·A·················· | ···················· | ···················· | ···················· | ··A················· |
| F | 5918 hCoV-19/Ireland/D-NVRL-70IRL97963/2020\|EPI_ISL_500 | ···················· | ···················· | ···················· | ···················· | ···················· | ···················· |
| F | 5919 hCoV-19/Ireland/WW-NVRL-70IRL98345/2020\|EPI_ISL_50 | ···················· | ·A·················· | ···················· | ···················· | ···················· | ··A················· |
| F | 5920 hCoV-19/Ireland/WD-NVRL-70IRL98816/2020\|EPI_ISL_50 | ···················· | ·A·················· | ···················· | ···················· | ···················· | ··A················· |
| F | 5921 hCoV-19/Ireland/D-NVRL-70IRL98924/2020\|EPI_ISL_500 | ···················· | ···················· | ···················· | ···················· | ···················· | ···················· |
| F | 5922 hCoV-19/Ireland/LK-NVRL-71IRL00996/2020\|EPI_ISL_50 | ···················· | ·A·················· | ···················· | ···················· | ···················· | ··A················· |
| F | 5923 hCoV-19/Ireland/D-NVRL-71IRL06434/2020\|EPI_ISL_500 | ···················· | ···················· | ···················· | ···················· | ···················· | ···················· |
| F | 5924 hCoV-19/Ireland/WD-NVRL-71IRL06455/2020\|EPI_ISL_50 | ···················· | ·A·················· | ···················· | ···················· | ···················· | ··A················· |
| F | 5925 hCoV-19/Ireland/WD-NVRL-71IRL06472/2020\|EPI_ISL_50 | ···················· | ·A·················· | ···················· | ···················· | ···················· | ··A················· |
| F | 5926 hCoV-19/Ireland/KK-NVRL-71IRL07034/2020\|EPI_ISL_50 | ···················· | ·A·················· | ···················· | ···················· | ···················· | ··A················· |
| F | 5927 hCoV-19/Ireland/LM-NVRL-71IRL07120/2020\|EPI_ISL_50 | ···················· | ·A·················· | ···················· | ···················· | ···················· | ··A················· |
| F | 5928 hCoV-19/Ireland/D-NVRL-71IRL07157/2020\|EPI_ISL_500 | ···················· | ·A·················· | ···················· | ···················· | ···················· | ··A················· |
| F | 5929 hCoV-19/Ireland/D-NVRL-71IRL07158/2020\|EPI_ISL_500 | ···················· | ···················· | ···················· | ···················· | ···················· | ···················· |
| F | 5930 hCoV-19/Ireland/D-NVRL-71IRL07177/2020\|EPI_ISL_500 | ···················· | ·A·················· | ···················· | ···················· | ···················· | ··A················· |
| F | 5931 hCoV-19/Ireland/D-NVRL-71IRL07206/2020\|EPI_ISL_500 | ···················· | ·A·················· | ···················· | ···················· | ···················· | ··A················· |
| F | 5932 hCoV-19/Ireland/D-NVRL-71IRL07248/2020\|EPI_ISL_500 | ···················· | ·A·················· | ···················· | ···················· | ···················· | ··N················· |
| F | 5933 hCoV-19/Australia/NSW2014/2020\|EPI_ISL_500600\|2020 | ···················· | ···················· | ···················· | ···················· | ···················· | ···················· |
| F | 5934 hCoV-19/Latvia/084/2020\|EPI_ISL_501915\|2020-06-16 | ···················· | ···················· | ···················· | ···················· | ···················· | ···················· |

| | | | | | | | |
|---|---|---|---|---|---|---|---|
| F | 5935 hCoV-19/Latvia/085/2020\|EPI_ISL_501922\|2020-06-16 | .................... | .................... | .................... | .................... | .................... | .................... |
| F | 5936 hCoV-19/Latvia/086/2020\|EPI_ISL_501929\|2020-07-09 | .................... | .................... | .................... | .................... | .................... | .................... |
| F | 5937 hCoV-19/Bangladesh/BCSIR-NILMRC-264/2020\|EPI_ISL_5 | .................... | .................... | .................... | .................... | .................... | .................... |
| F | 5938 hCoV-19/Bangladesh/BCSIR-NILMRC-284/2020\|EPI_ISL_5 | .................... | .................... | .................... | .................... | .................... | .................... |
| F | 5939 hCoV-19/Bangladesh/BCSIR-NILMRC-287/2020\|EPI_ISL_5 | .................... | .................... | .................... | .................... | .................... | .................... |
| F | 5940 hCoV-19/Bangladesh/BCSIR-NILMRC-292/2020\|EPI_ISL_5 | .................... | .................... | .................... | .................... | .................... | .................... |
| F | 5941 hCoV-19/Bangladesh/BCSIR-NILMRC-295/2020\|EPI_ISL_5 | .................... | .................... | .................... | .................... | .................... | .................... |
| F | 5942 hCoV-19/Bangladesh/BCSIR-NILMRC-317/2020\|EPI_ISL_5 | .................... | .................... | .................... | .................... | .................... | .................... |
| F | 5943 hCoV-19/Bangladesh/BCSIR-NILMRC-320/2020\|EPI_ISL_5 | .................... | .................... | .................... | .................... | .................... | .................... |
| F | 5944 hCoV-19/Bangladesh/BCSIR-NILMRC-330/2020\|EPI_ISL_5 | .................... | .................... | .................... | .................... | .................... | .................... |
| F | 5945 hCoV-19/Bangladesh/BCSIR-NILMRC-342/2020\|EPI_ISL_5 | .................... | .................... | .................... | .................... | .................... | .................... |
| F | 5946 hCoV-19/Bangladesh/BCSIR-NILMRC-343/2020\|EPI_ISL_5 | .................... | .A.................. | .................... | .................... | .................... | .................... |
| F | 5947 hCoV-19/Canada/AB-Z0001/2020\|EPI_ISL_504185\|2020-0 | .................... | .................... | .................... | .................... | .................... | .................... |
| F | 5948 hCoV-19/USA/MI-MDHHS-SC21124/2020\|EPI_ISL_507900\|2 | .................... | .................... | .................... | .................... | .................... | .................... |
| F | 5949 hCoV-19/USA/MI-MDHHS-SC21125/2020\|EPI_ISL_507901\|2 | .................... | .................... | .................... | .................... | .................... | .................... |
| F | 5950 hCoV-19/USA/MI-MDHHS-SC21126/2020\|EPI_ISL_507902\|2 | .................... | .................... | .................... | .................... | .................... | .................... |
| F | 5951 hCoV-19/USA/MI-MDHHS-SC21127/2020\|EPI_ISL_507903\|2 | .................... | .................... | .................... | .................... | .................... | .................... |
| F | 5952 hCoV-19/USA/MI-MDHHS-SC21128/2020\|EPI_ISL_507904\|2 | .................... | .................... | .................... | .................... | .................... | .................... |
| F | 5953 hCoV-19/USA/MI-MDHHS-SC21129/2020\|EPI_ISL_507905\|2 | .................... | .................... | .................... | .................... | .................... | .................... |
| F | 5954 hCoV-19/USA/MI-MDHHS-SC21130/2020\|EPI_ISL_507906\|2 | .................... | .................... | .................... | .................... | .................... | .................... |
| F | 5955 hCoV-19/USA/MI-MDHHS-SC21131/2020\|EPI_ISL_507907\|2 | .................... | .................... | .................... | .................... | .................... | .................... |
| F | 5956 hCoV-19/USA/MI-MDHHS-SC21132/2020\|EPI_ISL_507908\|2 | .................... | .................... | .................... | .................... | .................... | .................... |
| F | 5957 hCoV-19/USA/MI-MDHHS-SC21133/2020\|EPI_ISL_507909\|2 | .................... | .................... | .................... | .................... | .................... | .................... |
| F | 5958 hCoV-19/USA/MI-MDHHS-SC21134/2020\|EPI_ISL_507910\|2 | .................... | .................... | .................... | .................... | .................... | .................... |
| F | 5959 hCoV-19/USA/MI-MDHHS-SC21135/2020\|EPI_ISL_507911\|2 | .................... | .................... | .................... | .................... | .................... | .................... |
| F | 5960 hCoV-19/USA/MI-MDHHS-SC21136/2020\|EPI_ISL_507912\|2 | .................... | .................... | .................... | .................... | .................... | .................... |
| F | 5961 hCoV-19/USA/MI-MDHHS-SC21137/2020\|EPI_ISL_507913\|2 | .................... | .................... | .................... | .................... | .................... | .................... |
| F | 5962 hCoV-19/USA/MI-MDHHS-SC21138/2020\|EPI_ISL_507914\|2 | .................... | .................... | .................... | .................... | .................... | .................... |
| F | 5963 hCoV-19/USA/MI-MDHHS-SC21139/2020\|EPI_ISL_507915\|2 | .................... | .................... | .................... | .................... | .................... | .................... |
| F | 5964 hCoV-19/Portugal/PT0880/2020\|EPI_ISL_511139\|2020-0 | .................... | .................... | .................... | .................... | .................... | .................... |
| F | 5965 hCoV-19/Portugal/PT0881/2020\|EPI_ISL_511140\|2020-0 | .................... | .................... | .................... | .................... | .................... | .................... |
| F | 5966 hCoV-19/Portugal/PT0882/2020\|EPI_ISL_511141\|2020-0 | .................... | .................... | .................... | .................... | .................... | .................... |
| F | 5967 hCoV-19/Portugal/PT0886/2020\|EPI_ISL_511144\|2020-0 | .................... | .................... | .................... | .................... | .................... | .................... |
| F | 5968 hCoV-19/Portugal/PT0888/2020\|EPI_ISL_511146\|2020-0 | .................... | .................... | .................... | .................... | .................... | .................... |
| F | 5969 hCoV-19/Portugal/PT0889/2020\|EPI_ISL_511147\|2020-0 | .................... | .................... | .................... | .................... | .................... | .................... |
| F | 5970 hCoV-19/Portugal/PT0890/2020\|EPI_ISL_511148\|2020-0 | .................... | .................... | .................... | .................... | .................... | .................... |
| F | 5971 hCoV-19/Portugal/PT0891/2020\|EPI_ISL_511149\|2020-0 | .................... | .................... | .................... | .................... | .................... | .................... |
| F | 5972 hCoV-19/Portugal/PT0892/2020\|EPI_ISL_511150\|2020-0 | .................... | .................... | .................... | .................... | .................... | .................... |
| F | 5973 hCoV-19/Portugal/PT0893/2020\|EPI_ISL_511151\|2020-0 | .................... | .................... | .................... | .................... | .................... | .................... |
| F | 5974 hCoV-19/Portugal/PT0894/2020\|EPI_ISL_511152\|2020-0 | .................... | .................... | .................... | .................... | .................... | .................... |
| F | 5975 hCoV-19/Portugal/PT0897/2020\|EPI_ISL_511154\|2020-0 | .................... | .................... | .................... | .................... | .................... | .................... |
| F | 5976 hCoV-19/Portugal/PT0898/2020\|EPI_ISL_511155\|2020-0 | .................... | .................... | .................... | .................... | .................... | .................... |
| F | 5977 hCoV-19/Portugal/PT0899/2020\|EPI_ISL_511156\|2020-0 | .................... | .................... | .................... | .................... | .................... | .................... |
| F | 5978 hCoV-19/Portugal/PT0900/2020\|EPI_ISL_511157\|2020-0 | .................... | .................... | .................... | .................... | .................... | .................... |
| F | 5979 hCoV-19/Portugal/PT0901/2020\|EPI_ISL_511158\|2020-0 | .................... | .................... | .................... | .................... | .................... | .................... |
| F | 5980 hCoV-19/Portugal/PT0902/2020\|EPI_ISL_511159\|2020-0 | .................... | .................... | .................... | .................... | .................... | .................... |
| F | 5981 hCoV-19/Portugal/PT0903/2020\|EPI_ISL_511160\|2020-0 | .................... | .................... | .................... | .................... | .................... | .................... |
| F | 5982 hCoV-19/Portugal/PT0904/2020\|EPI_ISL_511161\|2020-0 | .................... | .................... | .................... | .................... | .................... | .................... |
| F | 5983 hCoV-19/Portugal/PT0905/2020\|EPI_ISL_511162\|2020-0 | .................... | .................... | .................... | .................... | .................... | .................... |
| F | 5984 hCoV-19/Portugal/PT0906/2020\|EPI_ISL_511163\|2020-0 | .................... | .................... | .................... | .................... | .................... | .................... |
| F | 5985 hCoV-19/Portugal/PT0907/2020\|EPI_ISL_511164\|2020-0 | .................... | .................... | .................... | .................... | .................... | .................... |
| F | 5986 hCoV-19/Portugal/PT0908/2020\|EPI_ISL_511165\|2020-0 | .................... | .................... | .................... | .................... | .................... | .................... |
| F | 5987 hCoV-19/Portugal/PT0909/2020\|EPI_ISL_511166\|2020-0 | .................... | .................... | .................... | .................... | .................... | .................... |
| F | 5988 hCoV-19/Portugal/PT0910/2020\|EPI_ISL_511167\|2020-0 | .................... | .................... | .................... | .................... | .................... | .................... |
| F | 5989 hCoV-19/Portugal/PT0911/2020\|EPI_ISL_511168\|2020-0 | .................... | .................... | .................... | .................... | .................... | .................... |
| F | 5990 hCoV-19/Portugal/PT0912/2020\|EPI_ISL_511169\|2020-0 | .................... | .A.................. | .................... | .................... | .................... | .................... |
| F | 5991 hCoV-19/Portugal/PT0913/2020\|EPI_ISL_511170\|2020-0 | .................... | .A.................. | .................... | .................... | .................... | .................... |
| F | 5992 hCoV-19/Portugal/PT0915/2020\|EPI_ISL_511172\|2020-0 | .................... | .A.................. | .................... | .................... | .................... | .................... |
| F | 5993 hCoV-19/Portugal/PT0916/2020\|EPI_ISL_511173\|2020-0 | .................... | .A.................. | .................... | .................... | .................... | .................... |
| F | 5994 hCoV-19/Portugal/PT0917/2020\|EPI_ISL_511174\|2020-0 | .................... | .A.................. | .................... | .................... | .................... | .................... |
| F | 5995 hCoV-19/Portugal/PT0918/2020\|EPI_ISL_511175\|2020-0 | .................... | .A.................. | .................... | .................... | .................... | .................... |
| F | 5996 hCoV-19/Portugal/PT0919/2020\|EPI_ISL_511176\|2020-0 | .................... | .................... | .................... | .................... | .................... | .................... |
| F | 5997 hCoV-19/Portugal/PT0920/2020\|EPI_ISL_511177\|2020-0 | .................... | .................... | .................... | .................... | .................... | .................... |
| F | 5998 hCoV-19/Portugal/PT1255/2020\|EPI_ISL_511216\|2020-0 | .................... | .................... | .................... | .................... | .................... | .................... |
| F | 5999 hCoV-19/Portugal/PT1256/2020\|EPI_ISL_511217\|2020-0 | .................... | .................... | .................... | .................... | .................... | .................... |
| F | 6000 hCoV-19/Portugal/PT1257/2020\|EPI_ISL_511218\|2020-0 | .................... | .................... | .................... | .................... | .................... | .................... |
| F | 6001 hCoV-19/Portugal/PT1258/2020\|EPI_ISL_511219\|2020-0 | .................... | .................... | .................... | .................... | .................... | .................... |
| F | 6002 hCoV-19/Portugal/PT1259/2020\|EPI_ISL_511220\|2020-0 | .................... | .................... | .................... | .................... | .................... | .................... |
| F | 6004 hCoV-19/Portugal/PT1261/2020\|EPI_ISL_511222\|2020-0 | .................... | .................... | .................... | .................... | .................... | .................... |
| F | 6005 hCoV-19/Portugal/PT1262/2020\|EPI_ISL_511223\|2020-0 | .................... | .................... | .................... | .................... | .................... | .................... |
| F | 6006 hCoV-19/Portugal/PT1263/2020\|EPI_ISL_511224\|2020-0 | .................... | .................... | .................... | .................... | .................... | .................... |
| F | 6007 hCoV-19/Portugal/PT1264/2020\|EPI_ISL_511225\|2020-0 | .................... | .................... | .................... | .................... | .................... | .................... |
| F | 6008 hCoV-19/Portugal/PT1265/2020\|EPI_ISL_511226\|2020-0 | .................... | .................... | .................... | .................... | .................... | .................... |
| F | 6009 hCoV-19/Portugal/PT1267/2020\|EPI_ISL_511228\|2020-0 | .................... | .................... | .................... | .................... | .................... | .................... |
| F | 6010 hCoV-19/Portugal/PT1268/2020\|EPI_ISL_511229\|2020-0 | .................... | .A.................. | .................... | .................... | .................... | .................... |
| F | 6011 hCoV-19/Portugal/PT1269/2020\|EPI_ISL_511230\|2020-0 | .................... | .A.................. | .................... | .................... | .................... | .................... |
| F | 6012 hCoV-19/Portugal/PT1270/2020\|EPI_ISL_511231\|2020-0 | .................... | .................... | .................... | .................... | .................... | .................... |
| F | 6013 hCoV-19/Portugal/PT1271/2020\|EPI_ISL_511232\|2020-0 | .................... | .................... | .................... | .................... | .................... | .................... |
| F | 6014 hCoV-19/Portugal/PT1272/2020\|EPI_ISL_511233\|2020-0 | .................... | .................... | .................... | .................... | .................... | .................... |
| F | 6015 hCoV-19/Portugal/PT1273/2020\|EPI_ISL_511234\|2020-0 | .................... | .................... | .................... | .................... | .................... | .................... |

| | | | | | | | |
|---|---|---|---|---|---|---|---|
| F | 6016 hCoV-19/Portugal/PT1274/2020\|EPI_ISL_511235\|2020-0 | ···················· | ···················· | ····················· | ···················· | ···················· | ···················· |
| F | 6017 hCoV-19/Portugal/PT1275/2020\|EPI_ISL_511236\|2020-0 | ···················· | ···················· | ····················· | ···················· | ···················· | ···················· |
| F | 6018 hCoV-19/Portugal/PT1276/2020\|EPI_ISL_511237\|2020-0 | ···················· | ···················· | ····················· | ···················· | ···················· | ···················· |
| F | 6019 hCoV-19/England/PORT-2D1534/2020\|EPI_ISL_512400\|20 | ···················· | ···T················ | ····················· | ···················· | ···················· | ···················· |
| F | 6020 hCoV-19/England/PORT-2D166E/2020\|EPI_ISL_512407\|20 | ···················· | ···················· | ····················· | ···················· | ···················· | ···················· |
| F | 6021 hCoV-19/England/PORT-2D2032/2020\|EPI_ISL_512409\|20 | ···················· | ···················· | ····················· | ···················· | ···················· | ···················· |
| F | 6022 hCoV-19/England/PORT-2D2041/2020\|EPI_ISL_512410\|20 | ···················· | ···················· | ····················· | ···················· | ···················· | ···················· |
| F | 6023 hCoV-19/England/PORT-2D206F/2020\|EPI_ISL_512411\|20 | ···················· | ···················· | ····················· | ···················· | ···················· | ···················· |
| F | 6024 hCoV-19/England/PORT-2D209C/2020\|EPI_ISL_512412\|20 | ···················· | ···T················ | ····················· | ···················· | ···················· | ···················· |
| F | 6025 hCoV-19/England/PORT-2D3E0A/2020\|EPI_ISL_512415\|20 | ···················· | ···················· | ····················· | ···················· | ···················· | ···················· |
| F | 6026 hCoV-19/England/PORT-2D3E37/2020\|EPI_ISL_512416\|20 | ···················· | ···················· | ····················· | ···················· | ···················· | ···················· |
| F | 6027 hCoV-19/England/PORT-2D3E82/2020\|EPI_ISL_512417\|20 | ···················· | ···················· | ····················· | ···················· | ···················· | ···················· |
| F | 6028 hCoV-19/Scotland/CVR3926/2020\|EPI_ISL_512470\|2020- | ···················· | ···················· | ····················· | ···················· | ···················· | ···················· |
| F | 6030 hCoV-19/Scotland/CVR3929/2020\|EPI_ISL_512472\|2020- | ···················· | ···················· | ····················· | ···················· | ···················· | ···················· |
| F | 6031 hCoV-19/Scotland/CVR3931/2020\|EPI_ISL_512474\|2020- | ···················· | ···················· | ····················· | ···················· | ···················· | ···················· |
| F | 6032 hCoV-19/Scotland/CVR3946/2020\|EPI_ISL_512476\|2020- | ···················· | ···················· | ····················· | ···················· | ···················· | ···················· |
| F | 6033 hCoV-19/Scotland/CVR3957/2020\|EPI_ISL_512477\|2020- | ···················· | ···················· | ····················· | ···················· | ···················· | ···················· |
| F | 6034 hCoV-19/Scotland/CVR3959/2020\|EPI_ISL_512478\|2020- | ···················· | ···················· | ····················· | ···················· | ···················· | ···················· |
| F | 6035 hCoV-19/Scotland/CVR3960/2020\|EPI_ISL_512479\|2020- | ···················· | ···················· | ····················· | ···················· | ···················· | ···················· |
| F | 6036 hCoV-19/Scotland/CVR3961/2020\|EPI_ISL_512480\|2020- | ···················· | ···················· | ····················· | ···················· | ···················· | ···················· |
| F | 6037 hCoV-19/Wales/PHWC-1695BA/2020\|EPI_ISL_512481\|2020 | ···················· | ···················· | ····················· | ···················· | ···················· | ···················· |
| F | 6039 hCoV-19/Wales/PHWC-1695D8/2020\|EPI_ISL_512483\|2020 | ···················· | ···················· | ····················· | ···················· | ···················· | ···················· |
| F | 6040 hCoV-19/Wales/PHWC-1695E7/2020\|EPI_ISL_512484\|2020 | ···················· | ·A·················· | ····················· | ···················· | ···················· | ··A················· |
| F | 6042 hCoV-19/Wales/PHWC-169602/2020\|EPI_ISL_512486\|2020 | ···················· | ···················· | ····················· | ···················· | ···················· | ···················· |
| F | 6043 hCoV-19/Wales/PHWC-169611/2020\|EPI_ISL_512487\|2020 | ···················· | ···················· | ····················· | ···················· | ···················· | ···················· |
| F | 6044 hCoV-19/Wales/PHWC-169620/2020\|EPI_ISL_512488\|2020 | ···················· | ···················· | ····················· | ···················· | ···················· | ···················· |
| F | 6045 hCoV-19/Wales/PHWC-16963F/2020\|EPI_ISL_512489\|2020 | ···················· | ···················· | ····················· | ···················· | ···················· | ···················· |
| F | 6046 hCoV-19/Wales/PHWC-16964E/2020\|EPI_ISL_512490\|2020 | ···················· | ···················· | ····················· | ···················· | ···················· | ···················· |
| F | 6047 hCoV-19/Wales/PHWC-16965D/2020\|EPI_ISL_512491\|2020 | ···················· | ···················· | ····················· | ···················· | ···················· | ···················· |
| F | 6049 hCoV-19/Wales/PHWC-16967B/2020\|EPI_ISL_512493\|2020 | ···················· | ···················· | ····················· | ···················· | ···················· | ···················· |
| F | 6050 hCoV-19/Wales/PHWC-16968A/2020\|EPI_ISL_512494\|2020 | ···················· | ···················· | ····················· | ···················· | ···················· | ···················· |
| F | 6051 hCoV-19/Wales/PHWC-169699/2020\|EPI_ISL_512495\|2020 | ···················· | ···················· | ····················· | ···················· | ···················· | ···················· |
| F | 6052 hCoV-19/Wales/PHWC-1696A8/2020\|EPI_ISL_512496\|2020 | ···················· | ···················· | ····················· | ···················· | ···················· | ···················· |
| F | 6053 hCoV-19/Wales/PHWC-1696B7/2020\|EPI_ISL_512497\|2020 | ···················· | ···················· | ····················· | ···················· | ···················· | ···················· |
| F | 6054 hCoV-19/Wales/PHWC-1696C6/2020\|EPI_ISL_512498\|2020 | ···················· | ···················· | ····················· | ···················· | ···················· | ···················· |
| F | 6055 hCoV-19/Wales/PHWC-1696D5/2020\|EPI_ISL_512499\|2020 | ···················· | ···················· | ····················· | ···················· | ···················· | ···················· |
| F | 6056 hCoV-19/USA/FL-BPHL-0741/2020\|EPI_ISL_512547\|2020- | ···················· | ···················· | ····················· | ···················· | ···················· | ···················· |
| F | 6057 hCoV-19/USA/FL-BPHL-0743/2020\|EPI_ISL_512549\|2020- | ···················· | ···················· | ····················· | ···················· | ···················· | ···················· |
| F | 6059 hCoV-19/USA/FL-BPHL-0746/2020\|EPI_ISL_512552\|2020- | ···················· | ···················· | ····················· | ···················· | ···················· | ···················· |
| F | 6061 hCoV-19/USA/FL-BPHL-0749/2020\|EPI_ISL_512555\|2020- | ···················· | ···················· | ····················· | ···················· | ···················· | ···················· |
| F | 6062 hCoV-19/USA/FL-BPHL-0750/2020\|EPI_ISL_512556\|2020- | ···················· | ···················· | ····················· | ···················· | ···················· | ···················· |
| F | 6063 hCoV-19/USA/FL-BPHL-0751/2020\|EPI_ISL_512557\|2020- | ···················· | ···················· | ····················· | ···················· | ···················· | ···················· |
| F | 6064 hCoV-19/USA/FL-BPHL-0754/2020\|EPI_ISL_512560\|2020- | ···················· | ···················· | ····················· | ···················· | ···················· | ···················· |
| F | 6065 hCoV-19/USA/FL-BPHL-0755/2020\|EPI_ISL_512561\|2020- | ···················· | ···················· | ····················· | ···················· | ···················· | ···················· |
| F | 6066 hCoV-19/USA/FL-BPHL-0756/2020\|EPI_ISL_512562\|2020- | ···················· | ···················· | ····················· | ···················· | ···················· | ···················· |
| F | 6067 hCoV-19/USA/FL-BPHL-0757/2020\|EPI_ISL_512563\|2020- | ···················· | ···················· | ····················· | ···················· | ···················· | ···················· |
| F | 6068 hCoV-19/USA/FL-BPHL-0758/2020\|EPI_ISL_512564\|2020- | ···················· | ···················· | ····················· | ···················· | ···················· | ···················· |
| F | 6069 hCoV-19/USA/FL-BPHL-0759/2020\|EPI_ISL_512565\|2020- | ···················· | ···················· | ····················· | ···················· | ···················· | ···················· |
| F | 6070 hCoV-19/USA/FL-BPHL-0761/2020\|EPI_ISL_512566\|2020- | ···················· | ···················· | ····················· | ···················· | ···················· | ···················· |
| F | 6071 hCoV-19/USA/FL-BPHL-0762/2020\|EPI_ISL_512567\|2020- | ···················· | ···················· | ····················· | ···················· | ···················· | ···················· |
| F | 6072 hCoV-19/USA/FL-BPHL-0763/2020\|EPI_ISL_512568\|2020- | ···················· | ···················· | ····················· | ···················· | ···················· | ···················· |
| F | 6073 hCoV-19/USA/UT-03002/2020\|EPI_ISL_513700\|2020-05-0 | ···················· | ···················· | ····················· | ···················· | ···················· | ···················· |
| F | 6074 hCoV-19/USA/UT-03006/2020\|EPI_ISL_513701\|2020-04-3 | ···················· | ···················· | ····················· | ···················· | ···················· | ···················· |
| F | 6075 hCoV-19/USA/UT-03014/2020\|EPI_ISL_513704\|2020-05-0 | ···················· | ···················· | ····················· | ···················· | ···················· | ···················· |
| F | 6076 hCoV-19/USA/CA-CZB-2157/2020\|EPI_ISL_513707\|2020-0 | ···················· | ···················· | ····················· | ···················· | ···················· | ···················· |
| F | 6077 hCoV-19/USA/CA-CZB-2158/2020\|EPI_ISL_513708\|2020-0 | ···················· | ···················· | ····················· | ···················· | ···················· | ···················· |
| F | 6078 hCoV-19/USA/CA-CZB-2159/2020\|EPI_ISL_513709\|2020-0 | ···················· | ···················· | ····················· | ···················· | ···················· | ···················· |
| F | 6079 hCoV-19/USA/CA-CZB-2160/2020\|EPI_ISL_513710\|2020-0 | ···················· | ···················· | ····················· | ···················· | ···················· | ···················· |
| F | 6080 hCoV-19/USA/CA-CZB-2161/2020\|EPI_ISL_513711\|2020-0 | ···················· | ···················· | ····················· | ···················· | ···················· | ···················· |
| F | 6081 hCoV-19/USA/CA-CZB-2162/2020\|EPI_ISL_513712\|2020-0 | ···················· | ···················· | ····················· | ···················· | ···················· | ···················· |
| F | 6082 hCoV-19/USA/CA-CZB-2163/2020\|EPI_ISL_513713\|2020-0 | ···················· | ···················· | ····················· | ···················· | ···················· | ···················· |
| F | 6083 hCoV-19/USA/CA-CZB-2164/2020\|EPI_ISL_513714\|2020-0 | ···················· | ···················· | ····················· | ···················· | ···················· | ···················· |
| F | 6084 hCoV-19/USA/CA-CZB-2165/2020\|EPI_ISL_513715\|2020-0 | ···············Y···· | ···················· | ····················· | ···················· | ···················· | ···················· |
| F | 6085 hCoV-19/USA/CA-CZB-2166/2020\|EPI_ISL_513716\|2020-0 | ···················· | ···················· | ····················· | ···················· | ···················· | ···················· |
| F | 6086 hCoV-19/USA/CA-CZB-2167/2020\|EPI_ISL_513717\|2020-0 | ···················· | ···················· | ····················· | ···················· | ···················· | ···················· |
| F | 6087 hCoV-19/USA/CA-CZB-2168/2020\|EPI_ISL_513718\|2020-0 | ···················· | ···················· | ····················· | ···················· | ···················· | ···················· |
| F | 6088 hCoV-19/USA/CA-CZB-2169/2020\|EPI_ISL_513719\|2020-0 | ···················· | ···················· | ····················· | ···················· | ···················· | ···················· |
| F | 6089 hCoV-19/USA/CA-CZB-2170/2020\|EPI_ISL_513720\|2020-0 | ···················· | ···················· | ····················· | ···················· | ···················· | ···················· |
| F | 6090 hCoV-19/USA/CA-CZB-2171/2020\|EPI_ISL_513721\|2020-0 | ···················· | ···················· | ····················· | ···················· | ···················· | ···················· |
| F | 6091 hCoV-19/USA/CA-CZB-2173/2020\|EPI_ISL_513723\|2020-0 | ···················· | ···················· | ····················· | ···················· | ···················· | ···················· |
| F | 6092 hCoV-19/USA/CA-CZB-2174/2020\|EPI_ISL_513724\|2020-0 | ···················· | ···················· | ····················· | ···················· | ···················· | ···················· |
| F | 6093 hCoV-19/USA/CA-CZB-2175/2020\|EPI_ISL_513725\|2020-0 | ···············Y···· | ·A·················· | ····················· | ···················· | ···················· | ···················· |
| F | 6094 hCoV-19/USA/CA-CZB-2176/2020\|EPI_ISL_513726\|2020-0 | ···················· | ···················· | ····················· | ···················· | ···················· | ···················· |
| F | 6095 hCoV-19/USA/CA-CZB-2177/2020\|EPI_ISL_513727\|2020-0 | ···················· | ···················· | ····················· | ···················· | ···················· | ···················· |
| F | 6096 hCoV-19/USA/CA-CZB-2178/2020\|EPI_ISL_513728\|2020-0 | ···················· | ···················· | ····················· | ···················· | ···················· | ···················· |
| F | 6097 hCoV-19/USA/CA-CZB-2179/2020\|EPI_ISL_513729\|2020-0 | ···················· | ···················· | ····················· | ···················· | ···················· | ···················· |
| F | 6098 hCoV-19/USA/CA-CZB-2184/2020\|EPI_ISL_513734\|2020-0 | ···················· | ···················· | ····················· | ···················· | ···················· | ···················· |
| F | 6099 hCoV-19/USA/CA-CZB-2186/2020\|EPI_ISL_513735\|2020-0 | ···················· | ···················· | ····················· | ···················· | ···················· | ···················· |
| F | 6100 hCoV-19/USA/CA-CZB-2188/2020\|EPI_ISL_513737\|2020-0 | ···················· | ···················· | ····················· | ···················· | ···················· | ···················· |
| F | 6101 hCoV-19/USA/CA-CZB-2189/2020\|EPI_ISL_513738\|2020-0 | ···················· | ···················· | ····················· | ···················· | ···················· | ···················· |

| | | | | | | | |
|---|---|---|---|---|---|---|---|
| F | 6102 hCoV-19/USA/CA-CZB-2190/2020\|EPI_ISL_513739\|2020-0 | ···················· | ···················· | ···················· | ···················· | ···················· | ···················· |
| F | 6103 hCoV-19/USA/CA-CZB-2191/2020\|EPI_ISL_513740\|2020-0 | ···················· | ···················· | ···················· | ···················· | ···················· | ···················· |
| F | 6104 hCoV-19/USA/CA-CZB-2192/2020\|EPI_ISL_513741\|2020-0 | ···················· | ···················· | ···················· | ···················· | ···················· | ···················· |
| F | 6105 hCoV-19/USA/CA-CZB-2193/2020\|EPI_ISL_513742\|2020-0 | ···················· | ···················· | ···················· | ···················· | ···················· | ···················· |
| F | 6106 hCoV-19/USA/CA-CZB-2194/2020\|EPI_ISL_513743\|2020-0 | ···················· | ···················· | ···················· | ···················· | ···················· | ···················· |
| F | 6107 hCoV-19/USA/CA-CZB-2195/2020\|EPI_ISL_513744\|2020-0 | ···················· | ···················· | ···················· | ···················· | ···················· | ···················· |
| F | 6108 hCoV-19/USA/CA-CZB-2196/2020\|EPI_ISL_513745\|2020-0 | ···················· | ···················· | ···················· | ···················· | ···················· | ···················· |
| F | 6109 hCoV-19/USA/CA-CZB-2197/2020\|EPI_ISL_513746\|2020-0 | ···················· | ···················· | ···················· | ···················· | ···················· | ···················· |
| F | 6110 hCoV-19/USA/CA-CZB-2198/2020\|EPI_ISL_513747\|2020-0 | ···················· | ···················· | ···················· | ···················· | ···················· | ···················· |
| F | 6111 hCoV-19/USA/CA-CZB-2199/2020\|EPI_ISL_513748\|2020-0 | ···················· | ···················· | ···················· | ···················· | ···················· | ···················· |
| F | 6112 hCoV-19/USA/CA-CZB-2200/2020\|EPI_ISL_513749\|2020-0 | ···················· | ···················· | ···················· | ···················· | ···················· | ···················· |
| F | 6113 hCoV-19/USA/CA-CZB-2201/2020\|EPI_ISL_513750\|2020-0 | ···················· | ···················· | ···················· | ···················· | ···················· | ···················· |
| F | 6114 hCoV-19/USA/CA-CZB-2202/2020\|EPI_ISL_513751\|2020-0 | ···················· | ···················· | ···················· | ···················· | ···················· | ···················· |
| F | 6115 hCoV-19/USA/CA-CZB-2203/2020\|EPI_ISL_513752\|2020-0 | ···················· | ···················· | ···················· | ···················· | ···················· | ···················· |
| F | 6116 hCoV-19/USA/CA-CZB-2205/2020\|EPI_ISL_513753\|2020-0 | ········T··········· | ···················· | ···················· | ···················· | ···················· | ···················· |
| F | 6117 hCoV-19/USA/CA-CZB-2206/2020\|EPI_ISL_513754\|2020-0 | ···················· | ···················· | ···················· | ···················· | ···················· | ···················· |
| F | 6118 hCoV-19/USA/CA-CZB-2207/2020\|EPI_ISL_513755\|2020-0 | ···················· | ···················· | ···················· | ···················· | ···················· | ···················· |
| F | 6119 hCoV-19/USA/CA-CZB-2208/2020\|EPI_ISL_513756\|2020-0 | ···················· | ···················· | ···················· | ···················· | ···················· | ···················· |
| F | 6120 hCoV-19/USA/CA-CZB-2476/2020\|EPI_ISL_513878\|2020-0 | ···················· | ···················· | ···················· | ···················· | ···················· | ···················· |
| F | 6121 hCoV-19/USA/CA-CZB-2484/2020\|EPI_ISL_513879\|2020-0 | ···················· | ···················· | ···················· | ···················· | ···················· | ···················· |
| F | 6122 hCoV-19/USA/CA-CZB-2486/2020\|EPI_ISL_513880\|2020-0 | ···················· | ···················· | ···················· | ···················· | ···················· | ···················· |
| F | 6123 hCoV-19/USA/CA-CZB-2488/2020\|EPI_ISL_513882\|2020-0 | ···················· | ···················· | ···················· | ···················· | ···················· | ···················· |
| F | 6124 hCoV-19/USA/CA-CZB-2492/2020\|EPI_ISL_513883\|2020-0 | ···················· | ···················· | ···················· | ···················· | ···················· | ···················· |
| F | 6125 hCoV-19/USA/CA-CZB-2497/2020\|EPI_ISL_513884\|2020-0 | ···················· | ·A·················· | ···················· | ···················· | ···················· | ···················· |
| F | 6127 hCoV-19/USA/CA-CZB-2506/2020\|EPI_ISL_513891\|2020-0 | ···················· | ·A·················· | ···················· | ···················· | ···················· | ···················· |
| F | 6128 hCoV-19/USA/CA-CZB-2507/2020\|EPI_ISL_513892\|2020-0 | ···················· | ·A·················· | ···················· | ···················· | ···················· | ···················· |
| F | 6129 hCoV-19/USA/CA-CZB-2508/2020\|EPI_ISL_513893\|2020-0 | ···················· | ·A·················· | ···················· | ···················· | ···················· | ···················· |
| F | 6130 hCoV-19/USA/CA-CZB-2509/2020\|EPI_ISL_513894\|2020-0 | ···················· | ·A·················· | ···················· | ···················· | ···················· | ···················· |
| F | 6131 hCoV-19/USA/CA-CZB-2520/2020\|EPI_ISL_513895\|2020-0 | ···················· | ·A·················· | ···················· | ···················· | ···················· | ···················· |
| F | 6132 hCoV-19/USA/CA-CZB-2521/2020\|EPI_ISL_513896\|2020-0 | ···················· | ·A·················· | ···················· | ···················· | ···················· | ···················· |
| F | 6133 hCoV-19/USA/CA-CZB-2522/2020\|EPI_ISL_513897\|2020-0 | ···················· | ···················· | ···················· | ···················· | ···················· | ···················· |
| F | 6134 hCoV-19/USA/CA-CZB-2523/2020\|EPI_ISL_513898\|2020-0 | ···················· | ···················· | ···················· | ···················· | ···················· | ···················· |
| F | 6135 hCoV-19/USA/CA-CZB-2524/2020\|EPI_ISL_513899\|2020-0 | ···················· | ···················· | ···················· | ···················· | ···················· | ···················· |
| F | 6136 hCoV-19/USA/CA-CZB-2525/2020\|EPI_ISL_513900\|2020-0 | ···················· | ···················· | ···················· | ···················· | ···················· | ···················· |
| F | 6137 hCoV-19/USA/CA-CZB-2526/2020\|EPI_ISL_513901\|2020-0 | ···················· | ···················· | ···················· | ···················· | ···················· | ···················· |
| F | 6138 hCoV-19/USA/CA-CZB-2531/2020\|EPI_ISL_513902\|2020-0 | ···················· | ···················· | ···················· | ···················· | ···················· | ···················· |
| F | 6139 hCoV-19/USA/CA-CZB-2534/2020\|EPI_ISL_513903\|2020-0 | ···················· | ···················· | ···················· | ···················· | ···················· | ···················· |
| F | 6140 hCoV-19/USA/CA-CZB-2537/2020\|EPI_ISL_513905\|2020-0 | ···················· | ···················· | ···················· | ···················· | ···················· | ···················· |
| F | 6141 hCoV-19/USA/CA-CZB-2539/2020\|EPI_ISL_513907\|2020-0 | ···················· | ···················· | ···················· | ···················· | ···················· | ···················· |
| F | 6142 hCoV-19/USA/CA-CZB-2546/2020\|EPI_ISL_513908\|2020-0 | ···················· | ···················· | ···················· | ···················· | ···················· | ···················· |
| F | 6143 hCoV-19/USA/CA-CZB-2547/2020\|EPI_ISL_513909\|2020-0 | ···················· | ···················· | ···················· | ···················· | ···················· | ···················· |
| F | 6144 hCoV-19/USA/CA-CZB-2548/2020\|EPI_ISL_513910\|2020-0 | ···················· | ···················· | ···················· | ···················· | ···················· | ···················· |
| F | 6145 hCoV-19/Argentina/PAIS-G0001/2020\|EPI_ISL_499083\|2 | ···················· | ···················· | ···················· | ···················· | ···················· | ···················· |
| F | 6148 hCoV-19/Spain/PV-IBV-004242/2020\|EPI_ISL_500306\|20 | ···················· | ···················· | ···················· | ···················· | ···················· | ···················· |
| F | 6159 hCoV-19/Spain/RI-IBV-006028/2020\|EPI_ISL_500458\|20 | ···················· | ···················· | ···················· | ···················· | ···················· | ···················· |
| F | 6160 hCoV-19/USA/TX-QDX-219/2020\|EPI_ISL_500459\|2020-04 | ···················· | ···················· | ···················· | ···················· | ···················· | ···················· |
| F | 6161 hCoV-19/Brazil/PE-IAM08/2020\|EPI_ISL_500460\|2020-0 | ···················· | ···················· | ···················· | ···················· | ···················· | ···················· |
| F | 6162 hCoV-19/Brazil/PE-IAM10/2020\|EPI_ISL_500461\|2020-0 | ···················· | ···················· | ···················· | ···················· | ···················· | ···················· |
| F | 6163 hCoV-19/Brazil/PE-IAM103/2020\|EPI_ISL_500462\|2020- | ···················· | ···················· | ···················· | ···················· | ···················· | ···················· |
| F | 6164 hCoV-19/Brazil/PE-IAM138/2020\|EPI_ISL_500463\|2020- | ···················· | ···················· | ···················· | ···················· | ···················· | ···················· |
| F | 6166 hCoV-19/Brazil/PE-IAM221/2020\|EPI_ISL_500473\|2020- | ···················· | ···················· | ···················· | ···················· | ···················· | ···················· |
| F | 6167 hCoV-19/Brazil/PE-IAM238/2020\|EPI_ISL_500475\|2020- | ···················· | ···················· | ···················· | ···················· | ···················· | ···················· |
| F | 6168 hCoV-19/Brazil/PE-IAM30/2020\|EPI_ISL_500477\|2020-0 | ···················· | ···················· | ···················· | ···················· | ···················· | ···················· |
| F | 6169 hCoV-19/Brazil/PE-IAM311/2020\|EPI_ISL_500480\|2020- | ···················· | ···················· | ···················· | ···················· | ···················· | ···················· |
| F | 6170 hCoV-19/Brazil/PE-IAM39/2020\|EPI_ISL_500481\|2020-0 | ···················· | ···················· | ···················· | ···················· | ···················· | ···················· |
| F | 6171 hCoV-19/Brazil/PE-IAM48/2020\|EPI_ISL_500482\|2020-0 | ···················· | ···················· | ···················· | ···················· | ···················· | ···················· |
| F | 6172 hCoV-19/Brazil/PE-IAM67/2020\|EPI_ISL_500483\|2020-0 | ···················· | ···················· | ···················· | ···················· | ···················· | ···················· |
| F | 6173 hCoV-19/Brazil/PE-IAM84/2020\|EPI_ISL_500484\|2020-0 | ···················· | ···················· | ···················· | ···················· | ···················· | ···················· |
| F | 6174 hCoV-19/Brazil/PE-IAM87/2020\|EPI_ISL_500485\|2020-0 | ···················· | ···················· | ···················· | ···················· | ···················· | ···················· |
| F | 6175 hCoV-19/Brazil/PE-IAM89/2020\|EPI_ISL_500486\|2020-0 | ···················· | ···················· | ···················· | ···················· | ···················· | ···················· |
| F | 6176 hCoV-19/Scotland/GCVR-1707E7/2020\|EPI_ISL_501635\|2 | ···················· | ···················· | ···················· | ···················· | ···················· | ···················· |
| F | 6177 hCoV-19/Latvia/074/2020\|EPI_ISL_501808\|2020-07-06 | ···················· | ···················· | ···················· | ···················· | ···················· | ···················· |
| F | 6178 hCoV-19/Bangladesh/BCSIR-NILMRC-262/2020\|EPI_ISL_5 | ···················· | ···················· | ···················· | ···················· | ···················· | ···················· |
| F | 6179 hCoV-19/USA/MI-MDHHS-SC21140/2020\|EPI_ISL_507916\|2 | ···················· | ···················· | ···················· | ···················· | ···················· | ···················· |
| F | 6180 hCoV-19/USA/MI-MDHHS-SC21141/2020\|EPI_ISL_507917\|2 | ···················· | ···················· | ···················· | ···················· | ···················· | ···················· |
| F | 6181 hCoV-19/USA/MI-MDHHS-SC21143/2020\|EPI_ISL_507919\|2 | ···················· | ···················· | ···················· | ···················· | ···················· | ···················· |
| F | 6182 hCoV-19/USA/MI-MDHHS-SC21144/2020\|EPI_ISL_507920\|2 | ···················· | ···················· | ···················· | ···················· | ···················· | ···················· |
| F | 6183 hCoV-19/USA/MI-MDHHS-SC21145/2020\|EPI_ISL_507921\|2 | ···················· | ···················· | ···················· | ···················· | ···················· | ···················· |
| F | 6184 hCoV-19/USA/MI-MDHHS-SC21146/2020\|EPI_ISL_507922\|2 | ···················· | ···················· | ···················· | ···················· | ···················· | ···················· |
| F | 6185 hCoV-19/USA/MI-MDHHS-SC21147/2020\|EPI_ISL_507923\|2 | ···················· | ···················· | ···················· | ···················· | ···················· | ···················· |
| F | 6186 hCoV-19/USA/MI-MDHHS-SC21148/2020\|EPI_ISL_507924\|2 | ···················· | ···················· | ···················· | ···················· | ···················· | ···················· |
| F | 6187 hCoV-19/USA/MI-MDHHS-SC21149/2020\|EPI_ISL_507925\|2 | ···················· | ···················· | ···················· | ···················· | ···················· | ···················· |
| F | 6188 hCoV-19/USA/MI-MDHHS-SC21150/2020\|EPI_ISL_507926\|2 | ···················· | ···················· | ···················· | ···················· | ···················· | ···················· |
| F | 6189 hCoV-19/USA/MI-MDHHS-SC21151/2020\|EPI_ISL_507927\|2 | ···················· | ···················· | ···················· | ···················· | ···················· | ···················· |
| F | 6190 hCoV-19/USA/MI-MDHHS-SC21152/2020\|EPI_ISL_507928\|2 | ···················· | ···················· | ···················· | ···················· | ···················· | ···················· |
| F | 6191 hCoV-19/USA/MI-MDHHS-SC21154/2020\|EPI_ISL_507930\|2 | ···················· | ···················· | ···················· | ···················· | ···················· | ···················· |
| F | 6192 hCoV-19/USA/MI-MDHHS-SC21155/2020\|EPI_ISL_507931\|2 | ···················· | ···················· | ···················· | ···················· | ···················· | ···················· |
| F | 6193 hCoV-19/USA/MI-MDHHS-SC21156/2020\|EPI_ISL_507932\|2 | ···················· | ···················· | ···················· | ···················· | ···················· | ···················· |
| F | 6194 hCoV-19/USA/MI-MDHHS-SC21157/2020\|EPI_ISL_507933\|2 | ···················· | ···················· | ···················· | ···················· | ···················· | ···················· |
| F | 6195 hCoV-19/USA/MN-MDH-1345/2020\|EPI_ISL_507934\|2020-0 | ···················· | ···················· | ···················· | ···················· | ···················· | ···················· |

| | | | | | | | | | |
|---|---|---|---|---|---|---|---|---|---|
| F | 6196 | hCoV-19/USA/MN-MDH-1346/2020\|EPI_ISL_507935\|2020-0 | ···················· | ···················· | ····················· | ···················· | ···················· | ···················· | |
| F | 6197 | hCoV-19/USA/MN-MDH-1347/2020\|EPI_ISL_507936\|2020-0 | ···················· | ···················· | ····················· | ···················· | ···················· | ···················· | |
| F | 6198 | hCoV-19/USA/MN-MDH-1348/2020\|EPI_ISL_507937\|2020-0 | ···················· | ···················· | ····················· | ···················· | ···················· | ···················· | |
| F | 6199 | hCoV-19/USA/MN-MDH-1349/2020\|EPI_ISL_507938\|2020-0 | ···················· | ···················· | ····················· | ···················· | ···················· | ···················· | |
| F | 6200 | hCoV-19/USA/MN-MDH-1350/2020\|EPI_ISL_507939\|2020-0 | ···················· | ···················· | ····················· | ···················· | ···················· | ···················· | |
| F | 6201 | hCoV-19/USA/MN-MDH-1351/2020\|EPI_ISL_507940\|2020-0 | ···················· | ···················· | ····················· | ···················· | ···················· | ···················· | |
| F | 6202 | hCoV-19/USA/MN-MDH-1352/2020\|EPI_ISL_507941\|2020-0 | ···················· | ···················· | ····················· | ···················· | ···················· | ···················· | |
| F | 6203 | hCoV-19/USA/MN-MDH-1353/2020\|EPI_ISL_507942\|2020-0 | ···················· | ···················· | ····················· | ···················· | ···················· | ···················· | |
| F | 6204 | hCoV-19/USA/MN-MDH-1354/2020\|EPI_ISL_507943\|2020-0 | ···················· | ···················· | ····················· | ···················· | ···················· | ···················· | |
| F | 6205 | hCoV-19/USA/MN-MDH-1355/2020\|EPI_ISL_507944\|2020-0 | ···················· | ···················· | ····················· | ···················· | ···················· | ···················· | |
| F | 6206 | hCoV-19/USA/MN-MDH-1356/2020\|EPI_ISL_507945\|2020-0 | ···················· | ···················· | ····················· | ···················· | ···················· | ···················· | |
| F | 6207 | hCoV-19/USA/MN-MDH-1357/2020\|EPI_ISL_507946\|2020-0 | ···················· | ···················· | ····················· | ···················· | NNN················· | ···················· | 1 |
| F | 6208 | hCoV-19/USA/MN-MDH-1358/2020\|EPI_ISL_507947\|2020-0 | ···················· | ···················· | ····················· | ···················· | ···················· | ···················· | |
| F | 6209 | hCoV-19/USA/MN-MDH-1359/2020\|EPI_ISL_507948\|2020-0 | ···················· | ···················· | ····················· | ···················· | ···················· | ···················· | |
| F | 6210 | hCoV-19/USA/MN-MDH-1360/2020\|EPI_ISL_507949\|2020-0 | ···················· | ···················· | ····················· | ···················· | ···················· | ···················· | |
| F | 6211 | hCoV-19/USA/MN-MDH-1361/2020\|EPI_ISL_507950\|2020-0 | ···················· | ···················· | ····················· | ···················· | ···················· | ···················· | |
| F | 6212 | hCoV-19/USA/MN-MDH-1362/2020\|EPI_ISL_507951\|2020-0 | ···················· | ···················· | ····················· | ···················· | ···················· | ···················· | |
| F | 6213 | hCoV-19/USA/MN-MDH-1363/2020\|EPI_ISL_507952\|2020-0 | ···················· | ···················· | ····················· | ···················· | ···················· | ···················· | |
| F | 6214 | hCoV-19/USA/MN-MDH-1365/2020\|EPI_ISL_507953\|2020-0 | ···················· | ···················· | ····················· | ···················· | ···················· | ···················· | |
| F | 6215 | hCoV-19/USA/MN-MDH-1366/2020\|EPI_ISL_507954\|2020-0 | ···················· | ···················· | ····················· | ···················· | ···················· | ···················· | |
| F | 6216 | hCoV-19/USA/MN-MDH-1367/2020\|EPI_ISL_507955\|2020-0 | ···················· | ···················· | ····················· | ···················· | ···················· | ···················· | |
| F | 6218 | hCoV-19/USA/MN-MDH-1369/2020\|EPI_ISL_507957\|2020-0 | ···················· | ·A·················· | ····················· | ···················· | ···················· | ···················· | |
| F | 6219 | hCoV-19/USA/MN-MDH-1370/2020\|EPI_ISL_507958\|2020-0 | ···················· | ·A·················· | ····················· | ···················· | ···················· | ···················· | |
| F | 6220 | hCoV-19/USA/MN-MDH-1371/2020\|EPI_ISL_507959\|2020-0 | ···················· | ···················· | ····················· | ···················· | ···················· | ···················· | |
| F | 6221 | hCoV-19/USA/MN-MDH-1372/2020\|EPI_ISL_507960\|2020-0 | ···················· | ·A·················· | ····················· | ···················· | ···················· | ···················· | |
| F | 6222 | hCoV-19/USA/MN-MDH-1373/2020\|EPI_ISL_507961\|2020-0 | ···················· | ···················· | ····················· | ···················· | ···················· | ···················· | |
| F | 6223 | hCoV-19/USA/MN-MDH-1374/2020\|EPI_ISL_507962\|2020-0 | ···················· | ···················· | ····················· | ···················· | ···················· | ···················· | |
| F | 6224 | hCoV-19/USA/MN-MDH-1375/2020\|EPI_ISL_507963\|2020-0 | ···················· | ···················· | ····················· | ···················· | ···················· | ···················· | |
| F | 6225 | hCoV-19/USA/MN-MDH-1376/2020\|EPI_ISL_507964\|2020-0 | ···················· | ···················· | ····················· | ···················· | ···················· | ···················· | |
| F | 6226 | hCoV-19/USA/MN-MDH-1377/2020\|EPI_ISL_507965\|2020-0 | ···················· | ···················· | ····················· | ···················· | ···················· | ···················· | |
| F | 6227 | hCoV-19/USA/MN-MDH-1378/2020\|EPI_ISL_507966\|2020-0 | ···················· | ···················· | ····················· | ···················· | ···················· | ···················· | |
| F | 6228 | hCoV-19/USA/MN-MDH-1379/2020\|EPI_ISL_507967\|2020-0 | ···················· | ···················· | ····················· | ···················· | ···················· | ···················· | |
| F | 6229 | hCoV-19/USA/MN-MDH-1380/2020\|EPI_ISL_507968\|2020-0 | ···················· | ···················· | ····················· | ···················· | ···················· | ···················· | |
| F | 6230 | hCoV-19/USA/MN-MDH-1381/2020\|EPI_ISL_507969\|2020-0 | ···················· | ···················· | ····················· | ···················· | ···················· | ···················· | |
| F | 6231 | hCoV-19/USA/MN-MDH-1382/2020\|EPI_ISL_507970\|2020-0 | ···················· | ···················· | ····················· | ···················· | ···················· | ···················· | |
| F | 6232 | hCoV-19/USA/MN-MDH-1383/2020\|EPI_ISL_507971\|2020-0 | ···················· | ···················· | ····················· | ···················· | ···················· | ···················· | |
| F | 6233 | hCoV-19/USA/MN-MDH-1384/2020\|EPI_ISL_507972\|2020-0 | ···················· | ···················· | ····················· | ···················· | ···················· | ···················· | |
| F | 6234 | hCoV-19/USA/MN-MDH-1386/2020\|EPI_ISL_507973\|2020-0 | ···················· | ···················· | ····················· | ···················· | ···················· | ···················· | |
| F | 6235 | hCoV-19/USA/MN-MDH-1387/2020\|EPI_ISL_507974\|2020-0 | ···················· | ···················· | ····················· | ···················· | ···················· | ···················· | |
| F | 6237 | hCoV-19/USA/MN-MDH-1389/2020\|EPI_ISL_507976\|2020-0 | ···················· | ···················· | ····················· | ···················· | ···················· | ···················· | |
| F | 6238 | hCoV-19/USA/MN-MDH-1390/2020\|EPI_ISL_507977\|2020-0 | ···················· | ···················· | ····················· | ···················· | ···················· | ···················· | |
| F | 6239 | hCoV-19/USA/MN-MDH-1391/2020\|EPI_ISL_507978\|2020-0 | ···················· | ···················· | ····················· | ···················· | ···················· | ···················· | |
| F | 6240 | hCoV-19/USA/MN-MDH-1392/2020\|EPI_ISL_507979\|2020-0 | ···················· | ···················· | ····················· | ···················· | ···················· | ···················· | |
| F | 6241 | hCoV-19/USA/MN-MDH-1393/2020\|EPI_ISL_507980\|2020-0 | ···················· | ···················· | ····················· | ···················· | ···················· | ···················· | |
| F | 6242 | hCoV-19/USA/MN-MDH-1394/2020\|EPI_ISL_507981\|2020-0 | ···················· | ···················· | ····················· | ···················· | ··N················· | ···················· | |
| F | 6243 | hCoV-19/USA/MN-MDH-1395/2020\|EPI_ISL_507982\|2020-0 | ···················· | ···················· | ····················· | ···················· | ···················· | ···················· | |
| F | 6244 | hCoV-19/USA/MN-MDH-1396/2020\|EPI_ISL_507983\|2020-0 | ···················· | ···················· | ····················· | ···················· | ···················· | ···················· | |
| F | 6245 | hCoV-19/USA/NM-UNM-00323/2020\|EPI_ISL_507986\|2020- | ···················· | ···················· | ····················· | ···················· | ···················· | ···················· | |
| F | 6246 | hCoV-19/USA/NM-UNM-00322/2020\|EPI_ISL_507987\|2020- | ···················· | ···················· | ····················· | ···················· | ···················· | ···················· | |
| F | 6247 | hCoV-19/USA/NM-UNM-00336/2020\|EPI_ISL_507991\|2020- | ···················· | ···················· | ····················· | ···················· | ···················· | ···················· | |
| F | 6248 | hCoV-19/USA/NM-UNM-00332/2020\|EPI_ISL_507995\|2020- | ···················· | ···················· | ····················· | ···················· | ···················· | ···················· | |
| F | 6249 | hCoV-19/Portugal/PT0841/2020\|EPI_ISL_511100\|2020-0 | ···················· | ·A·················· | ····················· | ···················· | ···················· | ···················· | |
| F | 6250 | hCoV-19/Portugal/PT0842/2020\|EPI_ISL_511101\|2020-0 | ···················· | ···················· | ····················· | ···················· | ···················· | ···················· | |
| F | 6251 | hCoV-19/Portugal/PT0843/2020\|EPI_ISL_511102\|2020-0 | ···················· | ···················· | ····················· | ···················· | ···················· | ···················· | |
| F | 6252 | hCoV-19/Portugal/PT0844/2020\|EPI_ISL_511103\|2020-0 | ···················· | ···················· | ····················· | ···················· | ···················· | ···················· | |
| F | 6253 | hCoV-19/Portugal/PT0845/2020\|EPI_ISL_511104\|2020-0 | ··T················· | ···················· | ····················· | ···················· | ···················· | ···················· | |
| F | 6254 | hCoV-19/Portugal/PT0848/2020\|EPI_ISL_511107\|2020-0 | ···················· | ···················· | ····················· | ···················· | ···················· | ···················· | |
| F | 6255 | hCoV-19/Portugal/PT0849/2020\|EPI_ISL_511108\|2020-0 | ···················· | ···················· | ····················· | ···················· | ···················· | ···················· | |
| F | 6256 | hCoV-19/Portugal/PT0850/2020\|EPI_ISL_511109\|2020-0 | ···················· | ···················· | ····················· | ···················· | ···················· | ···················· | |
| F | 6257 | hCoV-19/Portugal/PT0851/2020\|EPI_ISL_511110\|2020-0 | ···················· | ···················· | ····················· | ···················· | ···················· | ···················· | |
| F | 6258 | hCoV-19/Portugal/PT0852/2020\|EPI_ISL_511111\|2020-0 | ···················· | ···················· | ····················· | ···················· | ···················· | ···················· | |
| F | 6259 | hCoV-19/Portugal/PT0853/2020\|EPI_ISL_511112\|2020-0 | ···················· | ···················· | ····················· | ···················· | ···················· | ···················· | |
| F | 6260 | hCoV-19/Portugal/PT0854/2020\|EPI_ISL_511113\|2020-0 | ···················· | ···················· | ····················· | ···················· | ···················· | ···················· | |
| F | 6261 | hCoV-19/Portugal/PT0855/2020\|EPI_ISL_511114\|2020-0 | ···················· | ···················· | ····················· | ···················· | ···················· | ···················· | |
| F | 6262 | hCoV-19/Portugal/PT0856/2020\|EPI_ISL_511115\|2020-0 | ···················· | ···················· | ····················· | ···················· | ···················· | ···················· | |
| F | 6263 | hCoV-19/Portugal/PT0857/2020\|EPI_ISL_511116\|2020-0 | ···················· | ···················· | ····················· | ···················· | ···················· | ···················· | |
| F | 6264 | hCoV-19/Portugal/PT0858/2020\|EPI_ISL_511117\|2020-0 | ···················· | ···················· | ····················· | ···················· | ···················· | ···················· | |
| F | 6265 | hCoV-19/Portugal/PT0859/2020\|EPI_ISL_511118\|2020-0 | ···················· | ···················· | ····················· | ···················· | ···················· | ···················· | |
| F | 6266 | hCoV-19/Portugal/PT0860/2020\|EPI_ISL_511119\|2020-0 | ···················· | ···················· | ····················· | ···················· | ···················· | ···················· | |
| F | 6267 | hCoV-19/Portugal/PT0861/2020\|EPI_ISL_511120\|2020-0 | ···················· | ···················· | ····················· | ···················· | ···················· | ···················· | |
| F | 6268 | hCoV-19/Portugal/PT0862/2020\|EPI_ISL_511121\|2020-0 | ···················· | ···················· | ····················· | ···················· | ···················· | ···················· | |
| F | 6269 | hCoV-19/Portugal/PT0863/2020\|EPI_ISL_511122\|2020-0 | ···················· | ···················· | ····················· | ···················· | ···················· | ···················· | |
| F | 6270 | hCoV-19/Portugal/PT0864/2020\|EPI_ISL_511123\|2020-0 | ···················· | ···················· | ····················· | ···················· | ···················· | ···················· | |
| F | 6271 | hCoV-19/Portugal/PT0865/2020\|EPI_ISL_511124\|2020-0 | ···················· | ···················· | ····················· | ···················· | ···················· | ···················· | |
| F | 6272 | hCoV-19/Portugal/PT0866/2020\|EPI_ISL_511125\|2020-0 | ···················· | ···················· | ····················· | ···················· | ···················· | ···················· | |
| F | 6273 | hCoV-19/Portugal/PT0867/2020\|EPI_ISL_511126\|2020-0 | ···················· | ···················· | ····················· | ···················· | ···················· | ···················· | |
| F | 6274 | hCoV-19/Portugal/PT0868/2020\|EPI_ISL_511127\|2020-0 | ···················· | ···················· | ····················· | ···················· | ···················· | ···················· | |
| F | 6275 | hCoV-19/Portugal/PT0869/2020\|EPI_ISL_511128\|2020-0 | ···················· | ···················· | ····················· | ···················· | ···················· | ···················· | |
| F | 6276 | hCoV-19/Portugal/PT0870/2020\|EPI_ISL_511129\|2020-0 | ···················· | ···················· | ····················· | ···················· | ···················· | ···················· | |
| F | 6277 | hCoV-19/Portugal/PT0871/2020\|EPI_ISL_511130\|2020-0 | ···················· | ···················· | ····················· | ···················· | ···················· | ···················· | |

| | | | | | | | | | | | |
|---|---|---|---|---|---|---|---|---|---|---|---|
| F | 6278 | hCoV-19/Portugal/PT0872/2020 | EPI_ISL_511131 | 2020-0 | ···················· | ···················· | ···················· | ···················· | ···················· | ···················· | |
| F | 6279 | hCoV-19/Portugal/PT0873/2020 | EPI_ISL_511132 | 2020-0 | ···················· | ···················· | ···················· | ···················· | ···················· | ···················· | |
| F | 6280 | hCoV-19/Portugal/PT0874/2020 | EPI_ISL_511133 | 2020-0 | ···················· | ···················· | ···················· | ···················· | ···················· | ···················· | |
| F | 6281 | hCoV-19/Portugal/PT0875/2020 | EPI_ISL_511134 | 2020-0 | ···················· | ···················· | ···················· | ···················· | ···················· | ···················· | |
| F | 6282 | hCoV-19/Portugal/PT0876/2020 | EPI_ISL_511135 | 2020-0 | ···················· | ···················· | ···················· | ···················· | ···················· | ···················· | |
| F | 6283 | hCoV-19/Portugal/PT0877/2020 | EPI_ISL_511136 | 2020-0 | ···················· | ···················· | ···················· | ···················· | ···················· | ···················· | |
| F | 6284 | hCoV-19/Portugal/PT0878/2020 | EPI_ISL_511137 | 2020-0 | ···················· | ···················· | ···················· | ···················· | ···················· | ···················· | |
| F | 6285 | hCoV-19/Portugal/PT0879/2020 | EPI_ISL_511138 | 2020-0 | ···················· | ···················· | ···················· | ···················· | ···················· | ···················· | |
| F | 6286 | hCoV-19/Portugal/PT0887/2020 | EPI_ISL_511145 | 2020-0 | ···················· | ···················· | ···················· | ···················· | ···················· | ···················· | |
| F | 6287 | hCoV-19/Portugal/PT0923/2020 | EPI_ISL_511180 | 2020-0 | ···················· | ···················· | ···················· | ···················· | ···················· | ···················· | |
| F | 6288 | hCoV-19/Portugal/PT0924/2020 | EPI_ISL_511181 | 2020-0 | ···················· | ···················· | ···················· | ···················· | ···················· | ···················· | |
| F | 6289 | hCoV-19/Portugal/PT0925/2020 | EPI_ISL_511182 | 2020-0 | ···················· | ···················· | ···················· | ···················· | ···················· | ···················· | |
| F | 6290 | hCoV-19/Portugal/PT0926/2020 | EPI_ISL_511183 | 2020-0 | ···················· | ···················· | ···················· | ···················· | ···················· | ···················· | |
| F | 6291 | hCoV-19/Portugal/PT0927/2020 | EPI_ISL_511184 | 2020-0 | ···················· | ···················· | ···················· | ···················· | ···················· | ···················· | |
| F | 6292 | hCoV-19/Portugal/PT0928/2020 | EPI_ISL_511185 | 2020-0 | ···················· | ···················· | ···················· | ···················· | ···················· | ···················· | |
| F | 6293 | hCoV-19/Portugal/PT0929/2020 | EPI_ISL_511186 | 2020-0 | ···················· | ···················· | ···················· | ···················· | ···················· | ···················· | |
| F | 6294 | hCoV-19/Portugal/PT0930/2020 | EPI_ISL_511187 | 2020-0 | ···················· | ···················· | ···················· | ···················· | ···················· | ···················· | |
| F | 6295 | hCoV-19/Portugal/PT0931/2020 | EPI_ISL_511188 | 2020-0 | ···················· | ···················· | ···················· | ···················· | ···················· | ···················· | |
| F | 6296 | hCoV-19/Portugal/PT1229/2020 | EPI_ISL_511190 | 2020-0 | ···················· | ···················· | ···················· | ···················· | ···················· | ···················· | |
| F | 6297 | hCoV-19/Portugal/PT1230/2020 | EPI_ISL_511191 | 2020-0 | ···················· | ···················· | ···················· | ···················· | ···················· | ···················· | |
| F | 6298 | hCoV-19/Portugal/PT1231/2020 | EPI_ISL_511192 | 2020-0 | ···················· | ···················· | ···················· | ···················· | ···················· | ···················· | |
| F | 6299 | hCoV-19/Portugal/PT1232/2020 | EPI_ISL_511193 | 2020-0 | ···················· | ···················· | ···················· | ···················· | ···················· | ···················· | |
| F | 6301 | hCoV-19/Portugal/PT1234/2020 | EPI_ISL_511195 | 2020-0 | ···················· | ···················· | ···················· | ···················· | ···················· | ···················· | |
| F | 6302 | hCoV-19/Portugal/PT1235/2020 | EPI_ISL_511196 | 2020-0 | ···················· | ···················· | ···················· | ···················· | ···················· | ···················· | |
| F | 6303 | hCoV-19/Portugal/PT1236/2020 | EPI_ISL_511197 | 2020-0 | ···················· | ···················· | ···················· | ···················· | ···················· | ···················· | |
| F | 6304 | hCoV-19/Portugal/PT1237/2020 | EPI_ISL_511198 | 2020-0 | ···················· | ···················· | ···················· | ···················· | ···················· | ···················· | |
| F | 6305 | hCoV-19/Portugal/PT1238/2020 | EPI_ISL_511199 | 2020-0 | ···················· | ···················· | ···················· | ···················· | ···················· | ···················· | |
| F | 6306 | hCoV-19/Poland/1154951/2020 | EPI_ISL_512300 | 2020-06 | ···················· | ···················· | ···················· | ···················· | ···················· | ···················· | |
| F | 6307 | hCoV-19/Poland/1156085/2020 | EPI_ISL_512301 | 2020-06 | ···················· | ···················· | ···················· | ···················· | ···················· | ···················· | |
| F | 6308 | hCoV-19/Poland/1157220/2020 | EPI_ISL_512302 | 2020-06 | ···················· | ···················· | ···················· | ···················· | ···················· | ···················· | |
| F | 6309 | hCoV-19/Poland/1157821/2020 | EPI_ISL_512303 | 2020-06 | ···················· | ···················· | ···················· | ···················· | ···················· | ···················· | |
| F | 6310 | hCoV-19/England/PORT-2F5D50/2020 | EPI_ISL_512426 | 20 | ···················· | ···················· | ···················· | ···················· | ···················· | ···················· | |
| F | 6311 | hCoV-19/England/PORT-2F5DBA/2020 | EPI_ISL_512430 | 20 | ···················· | ···················· | ···················· | ···················· | ···················· | ···················· | |
| F | 6312 | hCoV-19/England/PORT-2F5DD8/2020 | EPI_ISL_512432 | 20 | ···················· | ···················· | ···················· | ···················· | ···················· | ···················· | |
| F | 6313 | hCoV-19/England/PORT-2F5DE7/2020 | EPI_ISL_512433 | 20 | ···················· | ···················· | ···················· | ···················· | ···················· | ···················· | |
| F | 6314 | hCoV-19/Scotland/CVR1288/2020 | EPI_ISL_512436 | 2020- | ···················· | ···················· | ···················· | ···················· | ···················· | ···················· | |
| F | 6316 | hCoV-19/Scotland/CVR1542/2020 | EPI_ISL_512439 | 2020- | ···················· | ···················· | ···················· | ···················· | ···················· | ···················· | |
| F | 6317 | hCoV-19/Scotland/CVR1546/2020 | EPI_ISL_512440 | 2020- | ···················· | ···················· | ···················· | ···················· | ···················· | ···················· | |
| F | 6318 | hCoV-19/Scotland/CVR1559/2020 | EPI_ISL_512443 | 2020- | ···················· | ···················· | ···················· | ···················· | ···················· | ···················· | |
| F | 6320 | hCoV-19/Scotland/CVR1666/2020 | EPI_ISL_512447 | 2020- | ···················· | ···················· | ···················· | ···················· | ···················· | ···················· | |
| F | 6326 | hCoV-19/Scotland/CVR1867/2020 | EPI_ISL_512455 | 2020- | ···················· | ···················· | ···················· | ···················· | ···················· | ···················· | |
| F | 6330 | hCoV-19/Scotland/CVR3908/2020 | EPI_ISL_512459 | 2020- | ···················· | ···················A | ···················· | ···················· | ···················· | ···················· | 1 |
| F | 6331 | hCoV-19/Scotland/CVR3910/2020 | EPI_ISL_512460 | 2020- | ···················· | ···················· | ···················· | ···················· | ···················· | ···················· | |
| F | 6332 | hCoV-19/Scotland/CVR3911/2020 | EPI_ISL_512461 | 2020- | ···················· | ···················A | ···················· | ···················· | ···················· | ···················· | 1 |
| F | 6334 | hCoV-19/Scotland/CVR3915/2020 | EPI_ISL_512463 | 2020- | ···················· | ···················· | ···················· | ···················· | ···················· | ···················· | |
| F | 6336 | hCoV-19/Scotland/CVR3921/2020 | EPI_ISL_512468 | 2020- | ···················· | ···················· | ···················· | ···················· | ···················· | ···················· | |
| F | 6339 | hCoV-19/DRC/6390/2020 | EPI_ISL_513615 | 2020-05-04 | ···················· | ···················· | ···················· | ···················· | ···················· | ···················· | |
| F | 6340 | hCoV-19/DRC/6395/2020 | EPI_ISL_513617 | 2020-05-07 | ···················· | ···················· | ···················· | ···················· | ···················· | ···················· | |
| F | 6341 | hCoV-19/DRC/6398/2020 | EPI_ISL_513618 | 2020-05-07 | ···················· | ···················· | ···················· | ···················· | ···················· | ···················· | |
| F | 6342 | hCoV-19/DRC/10733/2020 | EPI_ISL_513630 | 2020-05-23 | ···················· | ···················· | ···················· | ···················· | ···················· | ···················· | |
| F | 6343 | hCoV-19/USA/WA-UW-4607/2020 | EPI_ISL_513632 | 2020-03 | ···················· | ···················· | ···················· | ···················· | ···················· | ···················· | |
| F | 6344 | hCoV-19/USA/WA-UW-4660/2020 | EPI_ISL_513634 | 2020-04 | ···················· | ···················· | ···················· | ···················· | ···················· | ···················· | |
| F | 6345 | hCoV-19/USA/WA-UW-4664/2020 | EPI_ISL_513635 | 2020-04 | ···················· | ···················· | ···················· | ···················· | ···················· | ···················· | |
| F | 6346 | hCoV-19/USA/CA-CZB-2209/2020 | EPI_ISL_513757 | 2020-0 | ···················· | ···················· | ···················· | ···················· | ···················· | ···················· | |
| F | 6347 | hCoV-19/USA/CA-CZB-2210/2020 | EPI_ISL_513758 | 2020-0 | ···················· | ···················· | ···················· | ···················· | ···················· | ···················· | |
| F | 6348 | hCoV-19/USA/CA-CZB-2212/2020 | EPI_ISL_513760 | 2020-0 | ···················· | ···················· | ···················· | ···················· | ···················· | ···················· | |
| F | 6349 | hCoV-19/USA/CA-CZB-2213/2020 | EPI_ISL_513761 | 2020-0 | ···················· | ···················· | ···················· | ···················· | ···················· | ···················· | |
| F | 6350 | hCoV-19/USA/CA-CZB-2214/2020 | EPI_ISL_513762 | 2020-0 | ···················· | ···················· | ···················· | ···················· | ···················· | ···················· | |
| F | 6351 | hCoV-19/USA/CA-CZB-2218/2020 | EPI_ISL_513766 | 2020-0 | ···················· | ···················· | ···················· | ···················· | ···················· | ···················· | |
| F | 6352 | hCoV-19/USA/CA-CZB-2219/2020 | EPI_ISL_513767 | 2020-0 | ···················· | ·A·················· | ···················· | ···················· | ···················· | ···················· | |
| F | 6353 | hCoV-19/USA/CA-CZB-2220/2020 | EPI_ISL_513768 | 2020-0 | ···················· | ···················· | ···················· | ···················· | ···················· | ···················· | |
| F | 6354 | hCoV-19/USA/CA-CZB-2221/2020 | EPI_ISL_513769 | 2020-0 | ···················· | ···················· | ···················· | ···················· | ···················· | ···················· | |
| F | 6355 | hCoV-19/USA/CA-CZB-2222/2020 | EPI_ISL_513770 | 2020-0 | ···················· | ···················· | ···················· | ···················· | ···················· | ···················· | |
| F | 6356 | hCoV-19/USA/CA-CZB-2223/2020 | EPI_ISL_513771 | 2020-0 | ···················· | ···················· | ···················· | ···················· | ···················· | ···················· | |
| F | 6357 | hCoV-19/USA/CA-CZB-2224/2020 | EPI_ISL_513772 | 2020-0 | ···················· | ···················· | ···················· | ···················· | ···················· | ···················· | |
| F | 6358 | hCoV-19/USA/CA-CZB-2277/2020 | EPI_ISL_513773 | 2020-0 | ···················· | ···················· | ···················· | ···················· | ···················· | ···················· | |
| F | 6359 | hCoV-19/USA/CA-CZB-2278/2020 | EPI_ISL_513774 | 2020-0 | ···················· | ···················· | ···················· | ···················· | ···················· | ···················· | |
| F | 6360 | hCoV-19/USA/CA-CZB-2280/2020 | EPI_ISL_513775 | 2020-0 | ···················· | ···················· | ···················· | ···················· | ···················· | ···················· | |
| F | 6361 | hCoV-19/USA/CA-CZB-2281/2020 | EPI_ISL_513776 | 2020-0 | ···················· | ···················· | ···················· | ···················· | ···················· | ···················· | |
| F | 6362 | hCoV-19/USA/CA-CZB-2282/2020 | EPI_ISL_513777 | 2020-0 | ···················· | ···················· | ···················· | ···················· | ···················· | ···················· | |
| F | 6363 | hCoV-19/USA/CA-CZB-2284/2020 | EPI_ISL_513778 | 2020-0 | ···················· | ·A·················· | ···················· | ···················· | ···················· | ···················· | |
| F | 6364 | hCoV-19/USA/CA-CZB-2285/2020 | EPI_ISL_513779 | 2020-0 | ···················· | ·A·················· | ···················· | ···················· | ···················· | ···················· | |
| F | 6365 | hCoV-19/USA/CA-CZB-2286/2020 | EPI_ISL_513780 | 2020-0 | ···················· | ·A·················· | ···················· | ···················· | ···················· | ···················· | |
| F | 6366 | hCoV-19/USA/CA-CZB-2288/2020 | EPI_ISL_513781 | 2020-0 | ···················· | ···················· | ···················· | ···················· | ···················· | ·····C·············· | |
| F | 6367 | hCoV-19/USA/CA-CZB-2289/2020 | EPI_ISL_513782 | 2020-0 | ···················· | ·A·················· | ···················· | ···················· | ···················· | ···················· | |
| F | 6368 | hCoV-19/USA/CA-CZB-2292/2020 | EPI_ISL_513783 | 2020-0 | ···················· | ·A·················· | ···················· | ···················· | ···················· | ···················· | |
| F | 6369 | hCoV-19/USA/CA-CZB-2293/2020 | EPI_ISL_513784 | 2020-0 | ···················· | ·A·················· | ···················· | ···················· | ···················· | ···················· | |
| F | 6370 | hCoV-19/USA/CA-CZB-2295/2020 | EPI_ISL_513786 | 2020-0 | ···················· | ···················· | ···················· | ···················· | ···················· | ···················· | |
| F | 6371 | hCoV-19/USA/CA-CZB-2304/2020 | EPI_ISL_513788 | 2020-0 | ···················· | ···················· | ···················· | ···················· | ···················· | ···················· | |
| F | 6372 | hCoV-19/USA/CA-CZB-2339/2020 | EPI_ISL_513789 | 2020-0 | ···················· | ···················· | ···················· | ···················· | ···················· | ···················· | |

| | | | | | | | | | | |
|---|---|---|---|---|---|---|---|---|---|---|
| F | 6373 | hCoV-19/USA/CA-CZB-2340/2020\|EPI_ISL_513790\|2020-0 | ···················· | ···················· | ····················· | ···················· | ···················· | ···················· | | |
| F | 6374 | hCoV-19/USA/CA-CZB-2341/2020\|EPI_ISL_513791\|2020-0 | ···················· | ···················· | ····················· | ···················· | ···················· | ···················· | | |
| F | 6375 | hCoV-19/USA/CA-CZB-2342/2020\|EPI_ISL_513792\|2020-0 | ···················· | ···················· | ····················· | ···················· | ···················· | ···················· | | |
| F | 6376 | hCoV-19/USA/CA-CZB-2344/2020\|EPI_ISL_513793\|2020-0 | ···················· | ···················· | ····················· | ···················· | ···················· | ···················· | | |
| F | 6377 | hCoV-19/USA/CA-CZB-2345/2020\|EPI_ISL_513794\|2020-0 | ···················· | ···················· | ····················· | ···················· | ···················· | ···················· | | |
| F | 6378 | hCoV-19/USA/CA-CZB-2346/2020\|EPI_ISL_513795\|2020-0 | ···················· | ···················· | ····················· | ···················· | ···················· | ···················· | | |
| F | 6379 | hCoV-19/USA/CA-CZB-2347/2020\|EPI_ISL_513796\|2020-0 | ···················· | ···················· | ····················· | ···················· | ···················· | ···················· | | |
| F | 6380 | hCoV-19/USA/CA-CZB-2348/2020\|EPI_ISL_513797\|2020-0 | ···················· | ···················· | ····················· | ···················· | ···················· | ···················· | | |
| F | 6381 | hCoV-19/USA/CA-CZB-2350/2020\|EPI_ISL_513798\|2020-0 | ···················· | ···················· | ····················· | ···················· | ···················· | ···················· | | |
| F | 6382 | hCoV-19/USA/CA-CZB-2351/2020\|EPI_ISL_513799\|2020-0 | ···················· | ···················· | ····················· | ···················· | ···················· | ···················· | | |
| F | 6383 | hCoV-19/South Korea/KCDC2542/2020\|EPI_ISL_514945\|2 | ···················· | ···················· | ····················· | ···················· | ···················· | ···················· | | |
| F | 6384 | hCoV-19/South Korea/KCDC2544/2020\|EPI_ISL_514947\|2 | ···················· | ···················· | ····················· | ···················· | ···················· | ···················· | | |
| F | 6385 | hCoV-19/South Korea/KCDC2545/2020\|EPI_ISL_514948\|2 | ···················· | ···················· | ····················· | ···················· | ···················· | ···················· | | |
| F | 6386 | hCoV-19/South Korea/KCDC2546/2020\|EPI_ISL_514949\|2 | ···················· | ···················· | ····················· | ···················· | ···················· | ···················· | | |
| F | 6387 | hCoV-19/South Korea/KCDC2553/2020\|EPI_ISL_514956\|2 | ···················· | ···················· | ····················· | ···················· | ···················· | ···················· | | |
| F | 6388 | hCoV-19/South Korea/KCDC2554/2020\|EPI_ISL_514957\|2 | ···················· | ···················· | ····················· | ···················· | ···················· | ···················· | | |
| F | 6389 | hCoV-19/South Korea/KCDC2555/2020\|EPI_ISL_514958\|2 | ···················· | ···················· | ····················· | ···················· | ···················· | ···················· | | |
| F | 6390 | hCoV-19/South Korea/KCDC2556/2020\|EPI_ISL_514959\|2 | ···················· | ···················· | ····················· | ···················· | ···················· | ···················· | | |
| F | 6391 | hCoV-19/South Korea/KCDC2557/2020\|EPI_ISL_514960\|2 | ···················· | ···················· | ····················· | ···················· | ···················· | ···················· | | |
| F | 6392 | hCoV-19/South Korea/KCDC2558/2020\|EPI_ISL_514961\|2 | ···················· | ···················· | ····················· | ···················· | ···················· | ···················· | | |
| F | 6393 | hCoV-19/South Korea/KCDC2559/2020\|EPI_ISL_514962\|2 | ···················· | ···················· | ····················· | ···················· | ···················· | ···················· | | |
| F | 6394 | hCoV-19/South Korea/KCDC2560/2020\|EPI_ISL_514963\|2 | ···················· | ···················· | ····················· | ···················· | ···················· | ···················· | | |
| F | 6395 | hCoV-19/South Korea/KCDC2561/2020\|EPI_ISL_514964\|2 | ···················· | ···················· | ····················· | ···················· | ···················· | ···················· | | |
| F | 6396 | hCoV-19/South Korea/KCDC2562/2020\|EPI_ISL_514965\|2 | ···················· | ···················· | ····················· | ···················· | ···················· | ···················· | | |
| F | 6397 | hCoV-19/South Korea/KCDC2563/2020\|EPI_ISL_514966\|2 | ···················· | ···················· | ····················· | ···················· | ···················· | ···················· | | |
| F | 6398 | hCoV-19/Egypt/CUNCI-7I028/2020\|EPI_ISL_524426\|2020 | ···················· | ···················· | ····················· | ···················· | ···················· | ···················· | | |
| F | 6399 | hCoV-19/Egypt/CUNCI-7I026/2020\|EPI_ISL_524427\|2020 | ···················· | ···················· | ····················· | ···················· | ···················· | ···················· | | |
| F | 6400 | hCoV-19/USA/CA-SR-0840/2020\|EPI_ISL_524435\|2020-04 | ···················· | ···················· | ····················· | ···················· | ···················· | ···················· | | |
| F | 6401 | hCoV-19/India/MH-BJMC-838/2020\|EPI_ISL_496553\|2020 | ···················· | ···················· | ····················· | ···················· | ···················· | ···················· | | |
| F | 6402 | hCoV-19/India/MH-BJMC-965/2020\|EPI_ISL_496554\|2020 | ···················· | ···················· | ····················· | ···················· | ···················· | ···················· | | |
| F | 6403 | hCoV-19/India/MH-NCCS-NR048/2020\|EPI_ISL_496555\|20 | ···················· | ···················· | ····················· | ···················· | ···················· | ···················· | | |
| F | 6404 | hCoV-19/India/MH-NCCS-NR049/2020\|EPI_ISL_496556\|20 | ···················· | ···················· | ····················· | ···················· | ···················· | ···················· | | |
| F | 6405 | hCoV-19/India/MH-NCCS-NR051/2020\|EPI_ISL_496557\|20 | ···················· | ···················· | ····················· | ···················· | ···················· | ···················· | | |
| F | 6406 | hCoV-19/India/MH-NCCS-NR1290/2020\|EPI_ISL_496558\|2 | ···················· | ···················· | ····················· | ···················· | ···················· | ···················· | | |
| F | 6407 | hCoV-19/India/MH-NCCS-NR1323/2020\|EPI_ISL_496559\|2 | ···················· | ···················· | ····················· | ···················· | ···················· | ···················· | | |
| F | 6408 | hCoV-19/India/MH-NCCS-NR1709/2020\|EPI_ISL_496564\|2 | ···················· | ···················· | ····················· | ···················· | ···················· | ···················· | | |
| F | 6409 | hCoV-19/India/MH-NCCS-NR1715/2020\|EPI_ISL_496565\|2 | ···················· | ···················· | ····················· | ···················· | ···················· | ···················· | | |
| F | 6410 | hCoV-19/India/MH-NCCS-NR1885/2020\|EPI_ISL_496566\|2 | ···················· | ···················· | ····················· | ···················· | ···················· | ···················· | | |
| F | 6411 | hCoV-19/India/MH-NCCS-NR2086/2020\|EPI_ISL_496567\|2 | ···················· | ···················· | ····················· | ···················· | ···················· | ···················· | | |
| F | 6412 | hCoV-19/India/MH-NCCS-NR2099/2020\|EPI_ISL_496568\|2 | ···················· | ···················· | ····················· | ···················· | ···················· | ···················· | | |
| F | 6414 | hCoV-19/India/MH-NCCS-NR2109/2020\|EPI_ISL_496570\|2 | ···················· | ····A··············· | ····················· | ···················· | ···················· | ···················· | | |
| F | 6415 | hCoV-19/India/MH-NCCS-NR2122/2020\|EPI_ISL_496571\|2 | ···················· | ···················· | ····················· | ···················· | ···················· | ···················· | | |
| F | 6416 | hCoV-19/India/MH-NCCS-NR2245/2020\|EPI_ISL_496572\|2 | ···················· | ···················· | ····················· | ···················· | ···················· | ···················· | | |
| F | 6417 | hCoV-19/India/MH-NCCS-NR2246/2020\|EPI_ISL_496573\|2 | ···················· | ···················· | ····················· | ···················· | ···················· | ···················· | | |
| F | 6418 | hCoV-19/India/MH-NCCS-NR2249/2020\|EPI_ISL_496574\|2 | ···················· | ···················· | ····················· | ···················· | ···················· | ···················· | | |
| F | 6433 | hCoV-19/Spain/RI-IBV-005933/2020\|EPI_ISL_500369\|20 | ···················· | ···················· | ····················· | ···················· | ···················· | ···················· | | |
| F | 6435 | hCoV-19/Spain/RI-IBV-005940/2020\|EPI_ISL_500375\|20 | ···················· | ···················· | ····················· | ···················· | ···N················ | ···················· | | |
| F | 6438 | hCoV-19/Spain/RI-IBV-005952/2020\|EPI_ISL_500386\|20 | ···················· | ···················· | ····················· | ···················· | NNN················· | ···················· | | 1 |
| F | 6439 | hCoV-19/Spain/RI-IBV-005957/2020\|EPI_ISL_500391\|20 | ···················· | ···················· | ····················· | ···················· | ···················· | ···················· | | |
| F | 6440 | hCoV-19/Spain/RI-IBV-005962/2020\|EPI_ISL_500396\|20 | ···················· | ···················· | ····················· | ···················· | ···················· | ···················· | | |
| F | 6441 | hCoV-19/Spain/RI-IBV-005964/2020\|EPI_ISL_500398\|20 | ···················· | ···················· | ····················· | ···················· | ···················· | ···················· | | |
| F | 6442 | hCoV-19/Spain/RI-IBV-005965/2020\|EPI_ISL_500399\|20 | ···················· | ···················· | ····················· | ···················· | ···················· | ···················· | | |
| F | 6443 | hCoV-19/England/BRIS-130D78/2020\|EPI_ISL_501601\|20 | ···················· | ···················· | ····················· | ···················· | ···················· | ···················· | | |
| F | 6444 | hCoV-19/England/WSFT-25C15F2/2020\|EPI_ISL_501602\|2 | ···················· | ···················· | ····················· | ···················· | ···················· | ···················· | | |
| F | 6445 | hCoV-19/England/BRIS-130D87/2020\|EPI_ISL_501606\|20 | ···················· | ···················· | ····················· | ···················· | ···················· | ···················· | | |
| F | 6446 | hCoV-19/England/BRIS-130EFD/2020\|EPI_ISL_501607\|20 | ···················· | ···················· | ····················· | ···················· | ···················· | ···················· | | |
| F | 6447 | hCoV-19/England/BRIS-130FAF/2020\|EPI_ISL_501609\|20 | ··················T· | ···················· | ····················· | ···················· | ···················· | ···················· | 1 | |
| F | 6448 | hCoV-19/England/BRIS-130EB1/2020\|EPI_ISL_501611\|20 | ···················· | ···················· | ····················· | ···················· | ···················· | ···················· | | |
| F | 6449 | hCoV-19/England/WSFT-25C161D/2020\|EPI_ISL_501613\|2 | ···················· | ···················· | ····················· | ···················· | ···················· | ···················· | | |
| F | 6450 | hCoV-19/England/EKHU-260EF54/2020\|EPI_ISL_501614\|2 | ···················· | ···················· | ····················· | ···················· | ···················· | ···················· | | |
| F | 6451 | hCoV-19/England/EKHU-260F14E/2020\|EPI_ISL_501615\|2 | ···················· | ···················· | ····················· | ···················· | ···················· | ···················· | | |
| F | 6452 | hCoV-19/England/EKHU-260F18A/2020\|EPI_ISL_501616\|2 | ···················· | ···················· | ····················· | ···················· | ···················· | ···················· | | |
| F | 6453 | hCoV-19/England/EKHU-260F1B7/2020\|EPI_ISL_501617\|2 | ···················· | ···················· | ····················· | ···················· | ···················· | ···················· | | |
| F | 6454 | hCoV-19/England/EKHU-260F005/2020\|EPI_ISL_501618\|2 | ···················· | ···················· | ····················· | ···················· | ···················· | ···················· | | |
| F | 6455 | hCoV-19/England/EKHU-260EFBE/2020\|EPI_ISL_501620\|2 | ···················· | ···················· | ····················· | ···················· | ···················· | ···················· | | |
| F | 6456 | hCoV-19/England/EKHU-260EFFA/2020\|EPI_ISL_501621\|2 | ···················· | ···················· | ····················· | ···················· | ···················· | ···················· | | |
| F | 6457 | hCoV-19/England/EKHU-260F1D5/2020\|EPI_ISL_501622\|2 | ···················· | ···················· | ····················· | ···················· | ···················· | ···················· | | |
| F | 6458 | hCoV-19/England/EKHU-260F014/2020\|EPI_ISL_501623\|2 | ···················· | ···················· | ····················· | ···················· | ···················· | ···················· | | |
| F | 6459 | hCoV-19/England/EKHU-260EF81/2020\|EPI_ISL_501624\|2 | ···················· | ···················· | ····················· | ···················· | ···················· | ···················· | | |
| F | 6460 | hCoV-19/England/EKHU-260F023/2020\|EPI_ISL_501625\|2 | ···················· | ···················· | ····················· | ···················· | ···················· | ···················· | | |
| F | 6461 | hCoV-19/England/EKHU-260EF63/2020\|EPI_ISL_501626\|2 | ···················· | ···················· | ····················· | ···················· | ···················· | ···················· | | |
| F | 6462 | hCoV-19/England/EKHU-260F06F/2020\|EPI_ISL_501627\|2 | ···················· | ···················· | ····················· | ···················· | ···················· | ···················· | | |
| F | 6463 | hCoV-19/England/EKHU-260EFCD/2020\|EPI_ISL_501628\|2 | ···················· | ···················· | ····················· | ···················· | ···················· | ···················· | | |
| F | 6464 | hCoV-19/England/EKHU-260EF72/2020\|EPI_ISL_501629\|2 | ···················· | ···················· | ····················· | ···················· | ···················· | ···················· | | |
| F | 6465 | hCoV-19/Scotland/EDB2695/2020\|EPI_ISL_501632\|2020- | ···················· | ···················· | ····················· | ···················· | ···················· | ···················· | | |
| F | 6466 | hCoV-19/Scotland/GCVR-1708E4/2020\|EPI_ISL_501633\|2 | ···················· | ···················· | ····················· | ···················· | ···················· | ···················· | | |
| F | 6467 | hCoV-19/Scotland/GCVR-1712E5/2020\|EPI_ISL_501634\|2 | ···················· | ···················· | ····················· | ···················· | ···················· | ···················· | | |
| F | 6468 | hCoV-19/Portugal/PT0748/2020\|EPI_ISL_511007\|2020-0 | ···················· | ···················· | ····················· | ···················· | ···················· | ···················· | | |
| F | 6469 | hCoV-19/Portugal/PT0749/2020\|EPI_ISL_511008\|2020-0 | ···················· | ···················· | ····················· | ···················· | ···················· | ···················· | | |
| F | 6470 | hCoV-19/Portugal/PT0750/2020\|EPI_ISL_511009\|2020-0 | ···················· | ···················· | ····················· | ···················· | ···················· | ···················· | | |

| | | | | | | | | | | |
|---|---|---|---|---|---|---|---|---|---|---|
| F | 6471 | hCoV-19/Portugal/PT0751/2020\|EPI_ISL_511010\|2020-0 | .................... | .................... | .................... | .................... | .................... | .................... | | |
| F | 6472 | hCoV-19/Portugal/PT0752/2020\|EPI_ISL_511011\|2020-0 | .................... | .................... | .................... | .................... | .................... | .................... | | |
| F | 6473 | hCoV-19/Portugal/PT0753/2020\|EPI_ISL_511012\|2020-0 | .................... | .................... | .................... | .................... | .................... | .................... | | |
| F | 6474 | hCoV-19/Portugal/PT0755/2020\|EPI_ISL_511014\|2020-0 | .................... | .................... | .................... | .................... | .................... | .................... | | |
| F | 6475 | hCoV-19/Portugal/PT0757/2020\|EPI_ISL_511016\|2020-0 | .................... | .................... | .................... | .................... | .................... | .................... | | |
| F | 6476 | hCoV-19/Portugal/PT0758a/2020\|EPI_ISL_511017\|2020- | .................... | .................... | .................... | .................... | .................... | .................... | | |
| F | 6477 | hCoV-19/Portugal/PT0760/2020\|EPI_ISL_511019\|2020-0 | .................... | .................... | .................... | .................... | .................... | .................... | | |
| F | 6478 | hCoV-19/Portugal/PT0762/2020\|EPI_ISL_511021\|2020-0 | .................... | .................... | .................... | .................... | .................... | .................... | | |
| F | 6479 | hCoV-19/Portugal/PT0763/2020\|EPI_ISL_511022\|2020-0 | .................... | .................... | .................... | .................... | .................... | .................... | | |
| F | 6480 | hCoV-19/Portugal/PT0766/2020\|EPI_ISL_511025\|2020-0 | .................... | .................... | .................... | .................... | .................... | .................... | | |
| F | 6481 | hCoV-19/Portugal/PT0768/2020\|EPI_ISL_511027\|2020-0 | .................... | .................... | .................... | .................... | .................... | .................... | | |
| F | 6482 | hCoV-19/Portugal/PT0769a/2020\|EPI_ISL_511028\|2020- | .................... | .................... | .................... | .................... | .................... | .................... | | |
| F | 6483 | hCoV-19/Portugal/PT0770a/2020\|EPI_ISL_511029\|2020- | .................... | .................... | .................... | .................... | .................... | .................... | | |
| F | 6484 | hCoV-19/Portugal/PT0771/2020\|EPI_ISL_511030\|2020-0 | .................... | .................... | .................... | .................... | .................... | .................... | | |
| F | 6485 | hCoV-19/Portugal/PT0772/2020\|EPI_ISL_511031\|2020-0 | .................... | .................... | .................... | .................... | .................... | .................... | | |
| F | 6486 | hCoV-19/Portugal/PT0773/2020\|EPI_ISL_511032\|2020-0 | .................... | .................... | .................... | .................... | .................... | .................... | | |
| F | 6487 | hCoV-19/Portugal/PT0774/2020\|EPI_ISL_511033\|2020-0 | .................... | .................... | .................... | .................... | .................... | .................... | | |
| F | 6488 | hCoV-19/Portugal/PT0775/2020\|EPI_ISL_511034\|2020-0 | .................... | .................... | .................... | .................... | .................... | .................... | | |
| F | 6489 | hCoV-19/Portugal/PT0776/2020\|EPI_ISL_511035\|2020-0 | .................... | .................... | .................... | .................... | .................... | .................... | | |
| F | 6490 | hCoV-19/Portugal/PT0777a/2020\|EPI_ISL_511036\|2020- | .................... | .................... | .................... | .................... | .................... | .................... | | |
| F | 6491 | hCoV-19/Portugal/PT0778/2020\|EPI_ISL_511037\|2020-0 | .................... | .................... | .................... | .................... | .................... | .................... | | |
| F | 6492 | hCoV-19/Portugal/PT0780/2020\|EPI_ISL_511039\|2020-0 | .................... | .................... | .................... | .................... | .................... | .................... | | |
| F | 6493 | hCoV-19/Portugal/PT0781/2020\|EPI_ISL_511040\|2020-0 | .................... | .................... | .................... | .................... | .................... | .................... | | |
| F | 6494 | hCoV-19/Portugal/PT0782/2020\|EPI_ISL_511041\|2020-0 | .................... | .................... | .................... | .................... | .................... | .................... | | |
| F | 6495 | hCoV-19/Portugal/PT0783/2020\|EPI_ISL_511042\|2020-0 | .................... | .................... | .................... | .................... | .................... | .................... | | |
| F | 6496 | hCoV-19/Portugal/PT0784/2020\|EPI_ISL_511043\|2020-0 | .................... | .................... | .................... | .................... | .................... | .................... | | |
| F | 6497 | hCoV-19/Portugal/PT0786/2020\|EPI_ISL_511045\|2020-0 | .................... | .................... | .................... | .................... | .................... | .................... | | |
| F | 6498 | hCoV-19/Portugal/PT0787/2020\|EPI_ISL_511046\|2020-0 | .................... | .................... | .................... | .................... | .................... | .................... | | |
| F | 6499 | hCoV-19/Portugal/PT0788/2020\|EPI_ISL_511047\|2020-0 | .................... | .................... | .................... | .................... | .................... | .................... | | |
| F | 6500 | hCoV-19/Portugal/PT0791/2020\|EPI_ISL_511050\|2020-0 | .................... | .................... | .................... | .................... | .................... | .................... | | |
| F | 6501 | hCoV-19/Portugal/PT0793/2020\|EPI_ISL_511052\|2020-0 | .................... | .................... | .................... | .................... | .................... | .................... | | |
| F | 6502 | hCoV-19/Portugal/PT0794/2020\|EPI_ISL_511053\|2020-0 | .................... | .................... | .................... | .................... | .................... | .................... | | |
| F | 6503 | hCoV-19/Portugal/PT0795/2020\|EPI_ISL_511054\|2020-0 | .................... | .................... | .................... | .................... | .................... | .................... | | |
| F | 6504 | hCoV-19/Portugal/PT0796/2020\|EPI_ISL_511055\|2020-0 | .................... | .................... | .................... | .................... | .................... | .................... | | |
| F | 6505 | hCoV-19/Portugal/PT0797/2020\|EPI_ISL_511056\|2020-0 | .................... | .................... | .................... | .................... | .................... | .................... | | |
| F | 6506 | hCoV-19/Portugal/PT0798/2020\|EPI_ISL_511057\|2020-0 | .................... | .................... | .................... | .................... | .................... | .................... | | |
| F | 6507 | hCoV-19/Portugal/PT0799/2020\|EPI_ISL_511058\|2020-0 | .................... | .................... | .................... | .................... | .................... | .................... | | |
| F | 6508 | hCoV-19/Portugal/PT0800/2020\|EPI_ISL_511059\|2020-0 | .................... | .................... | .................... | .................... | .................... | .................... | | |
| F | 6509 | hCoV-19/England/EXET-13796E/2020\|EPI_ISL_512339\|20 | .................... | .................... | .................... | .................... | .................... | .................... | | |
| F | 6510 | hCoV-19/England/LIVE-A2158/2020\|EPI_ISL_512342\|202 | .................... | .................... | .................... | .................... | .................NNN | .................... | 1 | 1 |
| F | 6511 | hCoV-19/England/LIVE-A2765/2020\|EPI_ISL_512344\|202 | .................... | .................... | .................... | .................... | .................... | .................... | | |
| F | 6512 | hCoV-19/England/NORT-293950/2020\|EPI_ISL_512348\|20 | .................... | .................... | .................... | .................... | .................... | .................... | | |
| F | 6513 | hCoV-19/England/NORT-29397E/2020\|EPI_ISL_512349\|20 | .................... | .................... | .................... | .................... | .................... | .................... | | |
| F | 6514 | hCoV-19/England/NORT-2939E7/2020\|EPI_ISL_512350\|20 | .................... | .................... | .................... | .................... | .................... | .................... | | |
| F | 6515 | hCoV-19/England/NORT-293A7B/2020\|EPI_ISL_512351\|20 | .................... | .................... | .................... | .................... | .................... | .................... | | |
| F | 6516 | hCoV-19/England/NORT-293AD5/2020\|EPI_ISL_512353\|20 | .................... | .................... | .................... | .................... | .................... | .................... | | |
| F | 6517 | hCoV-19/England/NORT-293AE4/2020\|EPI_ISL_512354\|20 | .................... | .................... | .................... | .................... | .................... | .................... | | |
| F | 6518 | hCoV-19/England/NORT-293B4B/2020\|EPI_ISL_512356\|20 | .................... | .................... | .................... | .................... | .................... | .................... | | |
| F | 6519 | hCoV-19/England/NORT-293B78/2020\|EPI_ISL_512358\|20 | .................... | .................... | .................... | .................... | .................... | .................... | | |
| F | 6520 | hCoV-19/England/NORT-293B96/2020\|EPI_ISL_512359\|20 | .................... | .................... | .................... | .................... | .................... | .................... | | |
| F | 6521 | hCoV-19/England/NORT-293BE1/2020\|EPI_ISL_512360\|20 | .................... | .................... | .................... | .................... | .................... | .................... | | |
| F | 6522 | hCoV-19/England/NORT-293BF0/2020\|EPI_ISL_512361\|20 | .................... | .................... | .................... | .................... | .................... | .................... | | |
| F | 6523 | hCoV-19/England/NOTT-112E2E/2020\|EPI_ISL_512380\|20 | .................... | .................... | .................... | .................... | .................... | .................... | | |
| F | 6524 | hCoV-19/England/NOTT-112E5B/2020\|EPI_ISL_512381\|20 | .................... | .................... | .................... | .................... | .................... | .................... | | |
| F | 6525 | hCoV-19/England/NOTT-112E79/2020\|EPI_ISL_512382\|20 | .................... | .................... | .................... | .................... | .................... | .................... | | |
| F | 6526 | hCoV-19/England/PORT-2D1455/2020\|EPI_ISL_512389\|20 | .................... | .................... | .................... | .................... | .................... | .................... | | |
| F | 6527 | hCoV-19/DRC/4561/2020\|EPI_ISL_513600\|2020-04-30 | .................... | .................... | .................... | .................... | .................... | .................... | | |
| F | 6528 | hCoV-19/DRC/4562/2020\|EPI_ISL_513601\|2020-04-30 | .................... | .................... | .................... | .................... | .................... | .................... | | |
| F | 6529 | hCoV-19/DRC/4568/2020\|EPI_ISL_513602\|2020-04-30 | .................... | .................... | .................... | .................... | .................... | .................... | | |
| F | 6530 | hCoV-19/DRC/4613/2020\|EPI_ISL_513604\|2020-04-30 | .................... | .................... | .................... | .................... | .................... | .................... | | |
| F | 6531 | hCoV-19/DRC/4614/2020\|EPI_ISL_513605\|2020-04-30 | .................... | .................... | .................... | .................... | .................... | .................... | | |
| F | 6532 | hCoV-19/DRC/4631/2020\|EPI_ISL_513606\|2020-04-30 | .................... | .................... | .................... | .................... | .................... | .................... | | |
| F | 6533 | hCoV-19/DRC/4633/2020\|EPI_ISL_513607\|2020-04-30 | .................... | .................... | .................... | .................... | .................... | .................... | | |
| F | 6534 | hCoV-19/DRC/4642/2020\|EPI_ISL_513608\|2020-04-30 | .................... | .................... | .................... | .................... | .................... | .................... | | |
| F | 6535 | hCoV-19/DRC/4647/2020\|EPI_ISL_513609\|2020-04-30 | .................... | .................... | .................... | .................... | .................... | .................... | | |
| F | 6536 | hCoV-19/DRC/4653/2020\|EPI_ISL_513610\|2020-04-30 | .................... | .................... | .................... | .................... | .................... | .................... | | |
| F | 6537 | hCoV-19/DRC/6386/2020\|EPI_ISL_513613\|2020-05-07 | .................... | .................... | .................... | .................... | .................... | .................... | | |
| F | 6539 | hCoV-19/USA/UT-02866/2020\|EPI_ISL_513671\|2020-04-2 | .................... | .................... | .................... | .................... | .................... | .................... | | |
| F | 6540 | hCoV-19/USA/UT-02874/2020\|EPI_ISL_513675\|2020-04-2 | .................... | .................... | .................... | .................... | .................... | .................... | | |
| F | 6541 | hCoV-19/USA/UT-02878/2020\|EPI_ISL_513677\|2020-04-2 | .................... | .................... | .................... | .................... | .................... | .................... | | |
| F | 6542 | hCoV-19/USA/UT-02880/2020\|EPI_ISL_513678\|2020-04-2 | .................... | .................... | .................... | .................... | .................... | .................... | | |
| F | 6543 | hCoV-19/USA/UT-02888/2020\|EPI_ISL_513680\|2020-04-3 | .................... | .................... | .................... | .................... | .................... | .................... | | |
| F | 6544 | hCoV-19/USA/UT-02891/2020\|EPI_ISL_513683\|2020-04-2 | .................... | .................... | .................... | .................... | .................... | .................... | | |
| F | 6545 | hCoV-19/USA/UT-02893/2020\|EPI_ISL_513684\|2020-04-2 | .................... | .................... | .................... | .................... | .................... | .................... | | |
| F | 6546 | hCoV-19/USA/UT-02894/2020\|EPI_ISL_513685\|2020-04-3 | .................... | .................... | .................... | .................... | .................... | .................... | | |
| F | 6547 | hCoV-19/USA/UT-02895/2020\|EPI_ISL_513686\|2020-04-2 | .................... | .................... | .................... | .................... | .................... | .................... | | |
| F | 6548 | hCoV-19/USA/UT-02896/2020\|EPI_ISL_513687\|2020-04-3 | .................... | .................... | .................... | .................... | .................... | .................... | | |
| F | 6549 | hCoV-19/USA/UT-02897/2020\|EPI_ISL_513688\|2020-04-3 | .................... | .................... | .................... | .................... | .................... | .................... | | |
| F | 6550 | hCoV-19/USA/UT-02907/2020\|EPI_ISL_513690\|2020-04-3 | .................... | .................... | .................... | .................... | .................... | .................... | | |
| F | 6551 | hCoV-19/USA/UT-02908/2020\|EPI_ISL_513691\|2020-04-3 | .................... | .................... | .................... | .................... | .................... | .................... | | |

| | | | | | | | |
|---|---|---|---|---|---|---|---|
| F | 6552 hCoV-19/USA/UT-02917/2020\|EPI_ISL_513692\|2020-04-3 | ···················· | ···················· | ···················· | ···················· | ···················· | ···················· |
| F | 6553 hCoV-19/USA/UT-02940/2020\|EPI_ISL_513696\|2020-04-3 | ···················· | ···················· | ···················· | ···················· | ···················· | ···················· |
| F | 6554 hCoV-19/USA/UT-02941/2020\|EPI_ISL_513697\|2020-04-3 | ···················· | ···················· | ···················· | ···················· | ···················· | ···················· |
| F | 6555 hCoV-19/South Korea/KCDC2497/2020\|EPI_ISL_514900\|2 | ···················· | ···················· | ···················· | ···················· | ···················· | ···················· |
| F | 6556 hCoV-19/South Korea/KCDC2498/2020\|EPI_ISL_514901\|2 | ···················· | ···················· | ···················· | ···················· | ···················· | ···················· |
| F | 6557 hCoV-19/South Korea/KCDC2499/2020\|EPI_ISL_514902\|2 | ···················· | ···················· | ···················· | ···················· | ···················· | ···················· |
| F | 6558 hCoV-19/South Korea/KCDC2500/2020\|EPI_ISL_514903\|2 | ···················· | ···················· | ···················· | ···················· | ···················· | ···················· |
| F | 6559 hCoV-19/South Korea/KCDC2501/2020\|EPI_ISL_514904\|2 | ···················· | ···················· | ···················· | ···················· | ···················· | ···················· |
| F | 6560 hCoV-19/South Korea/KCDC2502/2020\|EPI_ISL_514905\|2 | ···················· | ···················· | ···················· | ···················· | ···················· | ···················· |
| F | 6561 hCoV-19/South Korea/KCDC2503/2020\|EPI_ISL_514906\|2 | ···················· | ···················· | ···················· | ···················· | ···················· | ···················· |
| F | 6562 hCoV-19/South Korea/KCDC2504/2020\|EPI_ISL_514907\|2 | ···················· | ···················· | ···················· | ···················· | ···················· | ···················· |
| F | 6563 hCoV-19/South Korea/KCDC2505/2020\|EPI_ISL_514908\|2 | ···················· | ···················· | ···················· | ···················· | ···················· | ···················· |
| F | 6564 hCoV-19/South Korea/KCDC2506/2020\|EPI_ISL_514909\|2 | ···················· | ···················· | ···················· | ···················· | ···················· | ···················· |
| F | 6565 hCoV-19/South Korea/KCDC2507/2020\|EPI_ISL_514910\|2 | ···················· | ···················· | ···················· | ···················· | ···················· | ···················· |
| F | 6566 hCoV-19/South Korea/KCDC2508/2020\|EPI_ISL_514911\|2 | ···················· | ···················· | ···················· | ···················· | ···················· | ···················· |
| F | 6567 hCoV-19/South Korea/KCDC2509/2020\|EPI_ISL_514912\|2 | ···················· | ···················· | ···················· | ···················· | ···················· | ···················· |
| F | 6568 hCoV-19/South Korea/KCDC2510/2020\|EPI_ISL_514913\|2 | ···················· | ···················· | ···················· | ···················· | ···················· | ···················· |
| F | 6569 hCoV-19/South Korea/KCDC2511/2020\|EPI_ISL_514914\|2 | ···················· | ···················· | ···················· | ···················· | ···················· | ···················· |
| F | 6570 hCoV-19/South Korea/KCDC2512/2020\|EPI_ISL_514915\|2 | ···················· | ···················· | ···················· | ···················· | ···················· | ···················· |
| F | 6571 hCoV-19/South Korea/KCDC2513/2020\|EPI_ISL_514916\|2 | ···················· | ···················· | ···················· | ···················· | ···················· | ···················· |
| F | 6572 hCoV-19/South Korea/KCDC2514/2020\|EPI_ISL_514917\|2 | ···················· | ···················· | ···················· | ···················· | ···················· | ···················· |
| F | 6573 hCoV-19/South Korea/KCDC2515/2020\|EPI_ISL_514918\|2 | ···················· | ···················· | ···················· | ···················· | ···················· | ···················· |
| F | 6574 hCoV-19/South Korea/KCDC2516/2020\|EPI_ISL_514919\|2 | ···················· | ···················· | ···················· | ···················· | ···················· | ···················· |
| F | 6575 hCoV-19/South Korea/KCDC2517/2020\|EPI_ISL_514920\|2 | ···················· | ···················· | ···················· | ···················· | ···················· | ···················· |
| F | 6576 hCoV-19/South Korea/KCDC2518/2020\|EPI_ISL_514921\|2 | ···················· | ···················· | ···················· | ···················· | ···················· | ···················· |
| F | 6577 hCoV-19/South Korea/KCDC2519/2020\|EPI_ISL_514922\|2 | ···················· | ···················· | ···················· | ···················· | ···················· | ···················· |
| F | 6578 hCoV-19/South Korea/KCDC2520/2020\|EPI_ISL_514923\|2 | ···················· | ···················· | ···················· | ···················· | ···················· | ···················· |
| F | 6579 hCoV-19/South Korea/KCDC2521/2020\|EPI_ISL_514924\|2 | ···················· | ···················· | ···················· | ···················· | ···················· | ···················· |
| F | 6580 hCoV-19/South Korea/KCDC2522/2020\|EPI_ISL_514925\|2 | ···················· | ···················· | ···················· | ···················· | ···················· | ···················· |
| F | 6581 hCoV-19/South Korea/KCDC2523/2020\|EPI_ISL_514926\|2 | ···················· | ···················· | ···················· | ···················· | ···················· | ···················· |
| F | 6582 hCoV-19/South Korea/KCDC2524/2020\|EPI_ISL_514927\|2 | ···················· | ···················· | ···················· | ···················· | ···················· | ···················· |
| F | 6583 hCoV-19/South Korea/KCDC2525/2020\|EPI_ISL_514928\|2 | ···················· | ···················· | ···················· | ···················· | ···················· | ···················· |
| F | 6584 hCoV-19/South Korea/KCDC2526/2020\|EPI_ISL_514929\|2 | ···················· | ···················· | ···················· | ···················· | ···················· | ···················· |
| F | 6585 hCoV-19/South Korea/KCDC2527/2020\|EPI_ISL_514930\|2 | ···················· | ···················· | ···················· | ···················· | ···················· | ···················· |
| F | 6586 hCoV-19/South Korea/KCDC2528/2020\|EPI_ISL_514931\|2 | ···················· | ···················· | ···················· | ···················· | ···················· | ···················· |
| F | 6587 hCoV-19/South Korea/KCDC2529/2020\|EPI_ISL_514932\|2 | ···················· | ···················· | ···················· | ···················· | ···················· | ···················· |
| F | 6588 hCoV-19/South Korea/KCDC2530/2020\|EPI_ISL_514933\|2 | ···················· | ···················· | ···················· | ···················· | ···················· | ···················· |
| F | 6589 hCoV-19/South Korea/KCDC2531/2020\|EPI_ISL_514934\|2 | ···················· | ···················· | ···················· | ···················· | ···················· | ···················· |
| F | 6590 hCoV-19/South Korea/KCDC2532/2020\|EPI_ISL_514935\|2 | ···················· | ···················· | ···················· | ···················· | ···················· | ···················· |
| F | 6591 hCoV-19/South Korea/KCDC2533/2020\|EPI_ISL_514936\|2 | ···················· | ···················· | ···················· | ···················· | ···················· | ···················· |
| F | 6592 hCoV-19/South Korea/KCDC2534/2020\|EPI_ISL_514937\|2 | ···················· | ···················· | ···················· | ···················· | ···················· | ···················· |
| F | 6593 hCoV-19/South Korea/KCDC2535/2020\|EPI_ISL_514938\|2 | ···················· | ···················· | ···················· | ···················· | ···················· | ···················· |
| F | 6594 hCoV-19/South Korea/KCDC2536/2020\|EPI_ISL_514939\|2 | ···················· | ···················· | ···················· | ···················· | ···················· | ···················· |
| F | 6595 hCoV-19/South Korea/KCDC2537/2020\|EPI_ISL_514940\|2 | ···················· | ···················· | ···················· | ···················· | ···················· | ···················· |
| F | 6596 hCoV-19/South Korea/KCDC2538/2020\|EPI_ISL_514941\|2 | ···················· | ···················· | ···················· | ···················· | ···················· | ···················· |
| F | 6597 hCoV-19/South Korea/KCDC2539/2020\|EPI_ISL_514942\|2 | ···················· | ···················· | ···················· | ···················· | ···················· | ···················· |
| F | 6598 hCoV-19/South Korea/KCDC2540/2020\|EPI_ISL_514943\|2 | ···················· | ···················· | ···················· | ···················· | ···················· | ···················· |
| F | 6599 hCoV-19/South Korea/KCDC2541/2020\|EPI_ISL_514944\|2 | ···················· | ···················· | ···················· | ···················· | ···················· | ···················· |
| F | 6600 hCoV-19/South Korea/KCDC2543/2020\|EPI_ISL_514946\|2 | ···················· | ···················· | ···················· | ···················· | ···················· | ···················· |
| F | 6601 hCoV-19/South Korea/KCDC2547/2020\|EPI_ISL_514950\|2 | ···················· | ···················· | ···················· | ···················· | ···················· | ···················· |
| F | 6602 hCoV-19/South Korea/KCDC2548/2020\|EPI_ISL_514951\|2 | ···················· | ···················· | ···················· | ···················· | ···················· | ···················· |
| F | 6603 hCoV-19/South Korea/KCDC2549/2020\|EPI_ISL_514952\|2 | ···················· | ···················· | ···················· | ···················· | ···················· | ···················· |
| F | 6604 hCoV-19/South Korea/KCDC2550/2020\|EPI_ISL_514953\|2 | ···················· | ···················· | ···················· | ···················· | ···················· | ···················· |
| F | 6605 hCoV-19/South Korea/KCDC2551/2020\|EPI_ISL_514954\|2 | ···················· | ···················· | ···················· | ···················· | ···················· | ···················· |
| F | 6606 hCoV-19/South Korea/KCDC2552/2020\|EPI_ISL_514955\|2 | ···················· | ···················· | ···················· | ···················· | ···················· | ···················· |
| F | 6607 hCoV-19/South Korea/KCDC2586/2020\|EPI_ISL_514989\|2 | ···················· | ···················· | ···················· | ···················· | ···················· | ···················· |
| F | 6608 hCoV-19/South Korea/KCDC2587/2020\|EPI_ISL_514990\|2 | ···················· | ···················· | ···················· | ···················· | ···················· | ···················· |
| F | 6609 hCoV-19/South Korea/KCDC2588/2020\|EPI_ISL_514991\|2 | ···················· | ···················· | ···················· | ···················· | ···················· | ···················· |
| F | 6610 hCoV-19/South Korea/KCDC2595/2020\|EPI_ISL_514998\|2 | ···················· | ···················· | ···················· | ···················· | ···················· | ···················· |
| F | 6611 hCoV-19/Brazil/RJ-DCVN3/2020\|EPI_ISL_529140\|2020-0 | ···················· | ···················· | ···················· | ···················· | ···················· | ···················· |
| F | 6612 hCoV-19/Egypt/CUNCI-HGC7I01/2020\|EPI_ISL_529141\|20 | ···················· | ···················· | ···················· | ···················· | ···················· | ···················· |
| F | 6613 hCoV-19/Egypt/CUNCI-HGC7I02/2020\|EPI_ISL_529142\|20 | ···················· | ···················· | ···················· | ···················· | ···················· | ···················· |
| F | 6614 hCoV-19/Egypt/CUNCI-HGC7I029/2020\|EPI_ISL_529143\|2 | ···················· | ···················· | ···················· | ···················· | ···················· | ···················· |
| F | 6615 hCoV-19/Egypt/CUNCI-HGC7I030/2020\|EPI_ISL_529144\|2 | ···················· | ···················· | ···················· | ···················· | ···················· | ···················· |
| F | 6616 hCoV-19/Egypt/CUNCI-HGC7I07/2020\|EPI_ISL_529145\|20 | ···················· | ······A············· | ···················· | ···················· | ···················· | ···················· |
| F | 6617 hCoV-19/USA/CA-ALSR-0183/2020\|EPI_ISL_529151\|2020- | ···················· | ···················· | ···················· | ···················· | ···················· | ···················· |
| F | 6618 hCoV-19/USA/CA-ALSR-0216/2020\|EPI_ISL_529152\|2020- | ···················· | ···················· | ···················· | ···················· | ···················· | ···················· |
| F | 6619 hCoV-19/USA/CA-ALSR-0231/2020\|EPI_ISL_529153\|2020- | ···················· | ·A·················· | ···················· | ···················· | ···················· | ···················· |
| F | 6620 hCoV-19/USA/CA-ALSR-1318/2020\|EPI_ISL_529154\|2020- | ···················· | ·A·················· | ···················· | ···················· | ···················· | ···················· |
| F | 6621 hCoV-19/USA/LA-ALSR-0142-NBG/2020\|EPI_ISL_529162\|2 | ···················· | ···················· | ···················· | ···················· | ···················· | ···················· |
| F | 6622 hCoV-19/USA/LA-ALSR-0165-NBG/2020\|EPI_ISL_529164\|2 | ···················· | ···················· | ···················· | ···················· | ···················· | ···················· |
| F | 6623 hCoV-19/USA/LA-ALSR-0171-NBG/2020\|EPI_ISL_529165\|2 | ···················· | ···················· | ···················· | ···················· | ···················· | ···················· |
| F | 6624 hCoV-19/USA/LA-ALSR-0172-NBG/2020\|EPI_ISL_529166\|2 | ···················· | ···················· | ···················· | ···················· | ···················· | ···················· |
| F | 6625 hCoV-19/USA/MD-MDH-0125/2020\|EPI_ISL_529167\|2020-0 | ···················· | ···················· | ···················· | ···················· | ···················· | ···················· |
| F | 6626 hCoV-19/USA/MD-MDH-0126/2020\|EPI_ISL_529168\|2020-0 | ···················· | ···················· | ···················· | ···················· | ···················· | ···················· |
| F | 6627 hCoV-19/USA/SC-DHEC-0014/2020\|EPI_ISL_529171\|2020- | ···················· | ···················· | ···················· | ···················· | ···················· | ···················· |
| F | 6628 hCoV-19/USA/SC-DHEC-0031/2020\|EPI_ISL_529173\|2020- | ···················· | ···················· | ···················· | ···················· | ···················· | ···················· |
| F | 6629 hCoV-19/USA/SC-DHEC-0032/2020\|EPI_ISL_529174\|2020- | ···················· | ···················· | ···················· | ···················· | ···················· | ···················· |
| F | 6630 hCoV-19/USA/SC-DHEC-0064/2020\|EPI_ISL_529175\|2020- | ···················· | ···················· | ···················· | ···················· | ···················· | ···················· |
| F | 6631 hCoV-19/USA/SC-DHEC-0074/2020\|EPI_ISL_529176\|2020- | ···················· | ··············C····· | ···················· | ···················· | ···················· | ···················· |

| | | | | | | | |
|---|---|---|---|---|---|---|---|
| F | 6632 hCoV-19/USA/SC-DHEC-0079/2020\|EPI_ISL_529178\|2020- | ···················· | ··············C····· | ···················· | ···················· | ···················· | ···················· |
| F | 6633 hCoV-19/USA/SC-DHEC-0116/2020\|EPI_ISL_529180\|2020- | ···················· | ········A··········· | ···················· | ···················· | ···················· | ···················· |
| F | 6634 hCoV-19/USA/SC-DHEC-0118/2020\|EPI_ISL_529181\|2020- | ···················· | ········A··········· | ···················· | ···················· | ···················· | ···················· |
| F | 6635 hCoV-19/USA/SC-DHEC-0119/2020\|EPI_ISL_529182\|2020- | ···················· | ········A··········· | ···················· | ···················· | ···················· | ···················· |
| F | 6636 hCoV-19/USA/SC-DHEC-0144/2020\|EPI_ISL_529183\|2020- | ···················· | ··············C····· | ···················· | ···················· | ···················· | ···················· |
| F | 6637 hCoV-19/USA/SC-DHEC-0156/2020\|EPI_ISL_529184\|2020- | ···················· | ···················· | ···················· | ···················· | ···················· | ···················· |
| F | 6638 hCoV-19/USA/SC-DHEC-0179/2020\|EPI_ISL_529185\|2020- | ···················· | ········A··········· | ···················· | ···················· | ···················· | ···················· |
| F | 6639 hCoV-19/USA/SC-DHEC-0180/2020\|EPI_ISL_529186\|2020- | ···················· | ········A··········· | ···················· | ···················· | ···················· | ···················· |
| F | 6640 hCoV-19/USA/SC-DHEC-0181/2020\|EPI_ISL_529187\|2020- | ···················· | ········A··········· | ···················· | ···················· | ···················· | ···················· |
| F | 6641 hCoV-19/USA/SC-DHEC-0182/2020\|EPI_ISL_529188\|2020- | ···················· | ········A··········· | ···················· | ···················· | ···················· | ···················· |
| F | 6642 hCoV-19/USA/SC-DHEC-0203/2020\|EPI_ISL_529190\|2020- | ···················· | ········A··········· | ···················· | ···················· | ···················· | ···················· |
| F | 6643 hCoV-19/USA/SC-DHEC-0204/2020\|EPI_ISL_529191\|2020- | ···················· | ········A··········· | ···················· | ···················· | ···················· | ···················· |
| F | 6644 hCoV-19/USA/SC-DHEC-0206/2020\|EPI_ISL_529192\|2020- | ···················· | ········A··········· | ···················· | ···················· | ···················· | ···················· |
| F | 6645 hCoV-19/USA/SC-DHEC-0207/2020\|EPI_ISL_529193\|2020- | ···················· | ········A··········· | ···················· | ···················· | ···················· | ···················· |
| F | 6646 hCoV-19/USA/SC-DHEC-0212/2020\|EPI_ISL_529194\|2020- | ···················· | ········A··········· | ···················· | ···················· | ···················· | ···················· |
| F | 6647 hCoV-19/USA/SC-DHEC-0233/2020\|EPI_ISL_529196\|2020- | ···················· | ········A··········· | ···················· | ···················· | ···················· | ···················· |
| F | 6648 hCoV-19/USA/SC-DHEC-0235/2020\|EPI_ISL_529197\|2020- | ···················· | ········A··········· | ···················· | ···················· | ···················· | ···················· |
| F | 6649 hCoV-19/USA/SC-DHEC-0236/2020\|EPI_ISL_529198\|2020- | ···················· | ········A··········· | ···················· | ···················· | ···················· | ···················· |
| F | 6650 hCoV-19/mink/Denmark/AD6-Farm1/2020\|EPI_ISL_626345 | ···················· | ···················· | ···················· | ···················· | ···················· | ···················· |
| F | 6651 hCoV-19/mink/Denmark/AD37-Farm3/2020\|EPI_ISL_62634 | ···················· | ···················· | ···················· | ···················· | ···················· | ···················· |
| F | 6652 hCoV-19/mink/Denmark/AL25-Farm1/2020\|EPI_ISL_62634 | ···················· | ···················· | ···················· | ···················· | ···················· | ···················· |
| F | 6653 hCoV-19/mink/Denmark/AL35-Farm3/2020\|EPI_ISL_62634 | ···················· | ···················· | ···················· | ···················· | ···················· | ···················· |
| F | 6654 hCoV-19/mink/Denmark/AL64-Farm1/2020\|EPI_ISL_62634 | ···················· | ···················· | ···················· | ···················· | ···················· | ···················· |
| F | 6655 hCoV-19/mink/Denmark/KL11-Farm1/2020\|EPI_ISL_62635 | ···················· | ···················· | ···················· | ···················· | ···················· | ···················· |
| F | 6656 hCoV-19/mink/Denmark/KL14-Farm1/2020\|EPI_ISL_62635 | ···················· | ···················· | ···················· | ···················· | ···················· | ···················· |
| F | 6657 hCoV-19/DRC/6875/2020\|EPI_ISL_496504\|2020-05-11 | ···················· | ···················· | ···················· | ···················· | ···················· | ···················· |
| F | 6658 hCoV-19/DRC/4418/2020\|EPI_ISL_496506\|2020-04-29 | ···················· | ···················· | ···················· | ···················· | ···················· | ···················· |
| F | 6659 hCoV-19/DRC/4430/2020\|EPI_ISL_496509\|2020-04-29 | ···················· | ···················· | ···················· | ···················· | ···················· | ···················· |
| F | 6660 hCoV-19/DRC/5186/2020\|EPI_ISL_496512\|2020-05-03 | ···················· | ···················· | ···················· | ···················· | ···················· | ···················· |
| F | 6661 hCoV-19/DRC/5377/2020\|EPI_ISL_496513\|2020-05-04 | ···················· | ···················· | ···················· | ···················· | ···················· | ···················· |
| F | 6662 hCoV-19/DRC/5996/2020\|EPI_ISL_496514\|2020-05-08 | ···················· | ···················· | ···················· | ···················· | ···················· | ···················· |
| F | 6663 hCoV-19/India/MH-AFMC-5286/2020\|EPI_ISL_496518\|202 | ···················· | ···················· | ···················· | ···················· | ···················· | ···················· |
| F | 6664 hCoV-19/India/MH-AFMC-5304/2020\|EPI_ISL_496519\|202 | ···················· | ···················· | ···················· | ···················· | ···················· | ···················· |
| F | 6665 hCoV-19/India/MH-AFMC-5473/2020\|EPI_ISL_496520\|202 | ···················· | ···················· | ···················· | ···················· | ···················· | ···················· |
| F | 6666 hCoV-19/India/MH-BJMC-1697/2020\|EPI_ISL_496521\|202 | ···················· | ···················· | ···················· | ···················· | ···················· | ···················· |
| F | 6667 hCoV-19/India/MH-BJMC-1769/2020\|EPI_ISL_496522\|202 | ···················· | ···················· | ···················· | ···················· | ···················· | ···················· |
| F | 6668 hCoV-19/India/MH-BJMC-6184/2020\|EPI_ISL_496523\|202 | ···················· | ···················· | ···················· | ···················· | ···················· | ···················· |
| F | 6669 hCoV-19/India/MH-NCCS-NR1205/2020\|EPI_ISL_496524\|2 | ···················· | ···················· | ···················· | ···················· | ···················· | ···················· |
| F | 6670 hCoV-19/India/MH-NCCS-NR1922/2020\|EPI_ISL_496525\|2 | ···················· | ···················· | ···················· | ···················· | ···················· | ···················· |
| F | 6671 hCoV-19/India/MH-NCCS-NR2257/2020\|EPI_ISL_496526\|2 | ···················· | ···················· | ···················· | ···················· | ···················· | ···················· |
| F | 6672 hCoV-19/India/MH-AFMC-5458/2020\|EPI_ISL_496529\|202 | ···················· | ···················· | ···················· | ···················· | ···················· | ···················· |
| F | 6673 hCoV-19/India/MH-BJMC-1089/2020\|EPI_ISL_496530\|202 | ···················· | ···················· | ···················· | ···················· | ···················· | ···················· |
| F | 6674 hCoV-19/India/MH-NCCS-NR2252/2020\|EPI_ISL_496531\|2 | ···················· | ···················· | ···················· | ···················· | ···················· | ···················· |
| F | 6675 hCoV-19/India/MH-NCCS-NR2309/2020\|EPI_ISL_496532\|2 | ···················· | ···················· | ···················· | ···················· | ···················· | ···················· |
| F | 6676 hCoV-19/India/MH-AFMC-5868/2020\|EPI_ISL_496533\|202 | ···················· | ···················· | ···················· | ···················· | ···················· | ···················· |
| F | 6677 hCoV-19/India/MH-BJMC-2035/2020\|EPI_ISL_496534\|202 | ···················· | ···················· | ···················· | ···················· | ···················· | ···················· |
| F | 6678 hCoV-19/India/MH-NCCS-NR2108/2020\|EPI_ISL_496535\|2 | ···················· | ···················· | ···················· | ···················· | ···················· | ···················· |
| F | 6679 hCoV-19/India/MH-AFMC-4954/2020\|EPI_ISL_496537\|202 | ···················· | ···················· | ···················· | ···················· | ···················· | ···················· |
| F | 6680 hCoV-19/India/MH-AFMC-5309/2020\|EPI_ISL_496538\|202 | ···················· | ···················· | ···················· | ···················· | ···················· | ···················· |
| F | 6681 hCoV-19/India/MH-AFMC-5318/2020\|EPI_ISL_496539\|202 | ···················· | ···················· | ···················· | ···················· | ···················· | ···················· |
| F | 6682 hCoV-19/India/MH-AFMC-5670/2020\|EPI_ISL_496540\|202 | ···················· | ···················· | ···················· | ···················· | ···················· | ···················· |
| F | 6683 hCoV-19/India/MH-AFMC-5688/2020\|EPI_ISL_496541\|202 | ···················· | ···················· | ···················· | ···················· | ···················· | ···················· |
| F | 6684 hCoV-19/India/MH-AFMC-5694/2020\|EPI_ISL_496542\|202 | ···················· | ···················· | ···················· | ···················· | ···················· | ···················· |
| F | 6685 hCoV-19/India/MH-AFMC-5695/2020\|EPI_ISL_496543\|202 | ···················· | ···················· | ···················· | ···················· | ···················· | ···················· |
| F | 6686 hCoV-19/India/MH-AFMC-5727/2020\|EPI_ISL_496544\|202 | ···················· | ···················· | ···················· | ···················· | ···················· | ···················· |
| F | 6687 hCoV-19/India/MH-AFMC-5807/2020\|EPI_ISL_496545\|202 | ···················· | ···················· | ···················· | ···················· | ···················· | ···················· |
| F | 6688 hCoV-19/India/MH-BJMC-1658/2020\|EPI_ISL_496546\|202 | ···················· | ···················· | ···················· | ···················· | ···················· | ···················· |
| F | 6689 hCoV-19/India/MH-BJMC-1675/2020\|EPI_ISL_496547\|202 | ···················· | ···················· | ···················· | ···················· | ···················· | ···················· |
| F | 6690 hCoV-19/India/MH-BJMC-1727/2020\|EPI_ISL_496548\|202 | ···················· | ···················· | ···················· | ···················· | ···················· | ···················· |
| F | 6691 hCoV-19/India/MH-BJMC-2030/2020\|EPI_ISL_496549\|202 | ···················· | ···················· | ···················· | ···················· | ···················· | ···················· |
| F | 6692 hCoV-19/India/MH-BJMC-2192/2020\|EPI_ISL_496550\|202 | ···················· | ···················· | ···················· | ···················· | ···················· | ···················· |
| F | 6693 hCoV-19/India/MH-BJMC-2442/2020\|EPI_ISL_496551\|202 | ···················· | ···················· | ···················· | ···················· | ···················· | ···················· |
| F | 6694 hCoV-19/India/MH-BJMC-6185/2020\|EPI_ISL_496552\|202 | ···················· | ···················· | ···················· | ···················· | ···················· | ···················· |
| F | 6695 hCoV-19/India/MH-NCCS-NR1326/2020\|EPI_ISL_496560\|2 | ···················· | ···················· | ···················· | ···················· | ···················· | ···················· |
| F | 6696 hCoV-19/India/MH-NCCS-NR1548/2020\|EPI_ISL_496561\|2 | ···················· | ···················· | ···················· | ···················· | ···················· | ···················· |
| F | 6697 hCoV-19/India/MH-NCCS-NR1553/2020\|EPI_ISL_496562\|2 | ···················· | ···················· | ···················· | ···················· | ···················· | ···················· |
| F | 6698 hCoV-19/India/MH-NCCS-NR1565/2020\|EPI_ISL_496563\|2 | ···················· | ···················· | ···················· | ···················· | ···················· | ···················· |
| F | 6699 hCoV-19/India/MH-NCCS-NR2254/2020\|EPI_ISL_496575\|2 | ···················· | ···················· | ···················· | ···················· | ···················· | ···················· |
| F | 6700 hCoV-19/India/MH-NCCS-NR2260/2020\|EPI_ISL_496576\|2 | ···················· | ·A·················· | ···················· | ···················· | ···················· | ··A················· |
| F | 6701 hCoV-19/India/MH-NCCS-NR2307/2020\|EPI_ISL_496577\|2 | ···················· | ···················· | ···················· | ···················· | ···················· | ···················· |
| F | 6702 hCoV-19/India/MH-NCCS-NR2328/2020\|EPI_ISL_496578\|2 | ···················· | ···················· | ···················· | ···················· | ···················· | ···················· |
| F | 6703 hCoV-19/India/MH-NCCS-NR2329/2020\|EPI_ISL_496579\|2 | ···················· | ···················· | ···················· | ···················· | ···················· | ···················· |
| F | 6704 hCoV-19/India/MH-NCCS-NR2347/2020\|EPI_ISL_496580\|2 | ···················· | ···················· | ···················· | ···················· | ···················· | ···················· |
| F | 6705 hCoV-19/India/MH-NCCS-NR2359/2020\|EPI_ISL_496581\|2 | ···················· | ···················· | ···················· | ···················· | ···················· | ···················· |
| F | 6706 hCoV-19/India/MH-NCCS-NR2513/2020\|EPI_ISL_496582\|2 | ···················· | ···················· | ···················· | ···················· | ···················· | ···················· |
| F | 6707 hCoV-19/India/MH-NCCS-NR2517/2020\|EPI_ISL_496583\|2 | ···················· | ···················· | ···················· | ···················· | ···················· | ···················· |
| F | 6708 hCoV-19/India/MH-NCCS-NR2523/2020\|EPI_ISL_496584\|2 | ···················· | ···················· | ···················· | ···················· | ···················· | ···················· |
| F | 6709 hCoV-19/India/MH-NCCS-NR2527/2020\|EPI_ISL_496585\|2 | ···················· | ···················· | ···················· | ···················· | ···················· | ···················· |
| F | 6710 hCoV-19/India/MH-NCCS-NR342/2020\|EPI_ISL_496586\|20 | ···················· | ···················· | ···················· | ···················· | ···················· | ···················· |
| F | 6711 hCoV-19/India/MH-NCCS-NR570/2020\|EPI_ISL_496587\|20 | ···················· | ···················· | ···················· | ···················· | ···················· | ···················· |

F 6712 hCoV-19/Hong Kong/HKU-200723-033/2020|EPI_ISL_4978 ···················· ···················· ···················· ···················· ···················· ····················
F 6713 hCoV-19/Hong Kong/HKU-200723-034/2020|EPI_ISL_4978 ···················· ···················· ···················· ···················· ···················· ····················
F 6714 hCoV-19/Hong Kong/HKU-200723-036/2020|EPI_ISL_4978 ···················· ·A·················· ···················· ···················· ···················· ··A·················
F 6715 hCoV-19/Hong Kong/HKU-200723-039/2020|EPI_ISL_4978 ···················· ···················· ···················· ···················· ···················· ····················
F 6716 hCoV-19/Hong Kong/HKU-200723-040/2020|EPI_ISL_4978 ···················· ···················· ···················· ···················· ···················· ····················
F 6717 hCoV-19/Hong Kong/HKU-200723-041/2020|EPI_ISL_4978 ···················· ···················· ···················· ···················· ···················· ····················
F 6718 hCoV-19/Hong Kong/HKU-200723-042/2020|EPI_ISL_4978 ···················· ···················· ···················· ···················· ···················· ····················
F 6719 hCoV-19/Hong Kong/HKU-200723-043/2020|EPI_ISL_4978 ···················· ···················· ···················· ···················· ···················· ····················
F 6720 hCoV-19/Hong Kong/HKU-200723-044/2020|EPI_ISL_4978 ···················· ···················· ···················· ···················· ···················· ····················
F 6721 hCoV-19/Hong Kong/HKU-200723-045/2020|EPI_ISL_4978 ···················· ···················· ···················· ···················· ···················· ····················
F 6722 hCoV-19/Hong Kong/HKU-200723-046/2020|EPI_ISL_4978 ···················· ···················· ···················· ···················· ···················· ····················
F 6723 hCoV-19/Hong Kong/HKU-200723-047/2020|EPI_ISL_4978 ···················· ···················· ···················· ···················· ···················· ····················
F 6724 hCoV-19/Hong Kong/HKU-200723-048/2020|EPI_ISL_4978 ···················· ···················· ···················· ···················· ···················· ····················
F 6725 hCoV-19/Hong Kong/HKU-200723-049/2020|EPI_ISL_4978 ···················· ···················· ···················· ···················· ···················· ····················
F 6726 hCoV-19/Hong Kong/HKU-200723-050/2020|EPI_ISL_4978 ···················· ···················· ···················· ···················· ···················· ····················
F 6727 hCoV-19/Hong Kong/HKU-200723-051/2020|EPI_ISL_4978 ···················· ···················· ···················· ···················· ···················· ····················
F 6728 hCoV-19/Hong Kong/HKU-200723-052/2020|EPI_ISL_4978 ···················· ···················· ···················· ···················· ···················· ····················
F 6729 hCoV-19/Hong Kong/HKU-200723-053/2020|EPI_ISL_4978 ···················· ···················· ···················· ···················· ···················· ····················
F 6730 hCoV-19/Hong Kong/HKU-200723-054/2020|EPI_ISL_4978 ···················· ···················· ···················· ···················· ···················· ····················
F 6731 hCoV-19/Hong Kong/HKU-200723-055/2020|EPI_ISL_4978 ···················· ···················· ···················· ···················· ···················· ····················
F 6732 hCoV-19/Hong Kong/HKU-200723-056/2020|EPI_ISL_4978 ···················· ···················· ···················· ···················· ···················· ····················
F 6733 hCoV-19/Hong Kong/HKU-200723-057/2020|EPI_ISL_4978 ···················· ···················· ···················· ···················· ···················· ····················
F 6734 hCoV-19/Hong Kong/HKU-200723-058/2020|EPI_ISL_4978 ···················· ···················· ···················· ···················· ···················· ····················
F 6735 hCoV-19/Hong Kong/HKU-200723-059/2020|EPI_ISL_4978 ···················· ···················· ···················· ···················· ···················· ····················
F 6736 hCoV-19/Hong Kong/HKU-200723-060/2020|EPI_ISL_4978 ···················· ···················· ···················· ···················· ···················· ····················
F 6737 hCoV-19/Hong Kong/HKU-200723-062/2020|EPI_ISL_4978 ···················· ···················· ···················· ···················· ···················· ····················
F 6738 hCoV-19/Hong Kong/HKU-200723-063/2020|EPI_ISL_4978 ···················· ···················· ···················· ···················· ···················· ····················
F 6739 hCoV-19/Hong Kong/HKU-200723-064/2020|EPI_ISL_4978 ···················· ···················· ···················· ···················· ···················· ····················
F 6740 hCoV-19/Hong Kong/HKU-200723-065/2020|EPI_ISL_4978 ···················· ···················· ···················· ···················· ···················· ····················
F 6741 hCoV-19/Hong Kong/HKU-200723-066/2020|EPI_ISL_4978 ···················· ···················· ···················· ···················· ···················· ····················
F 6742 hCoV-19/Hong Kong/HKU-200723-068/2020|EPI_ISL_4978 ···················· ···················· ···················· ···················· ···················· ····················
F 6743 hCoV-19/Hong Kong/HKU-200723-069/2020|EPI_ISL_4978 ···················· ···················· ···················· ···················· ···················· ····················
F 6744 hCoV-19/Hong Kong/HKU-200723-073/2020|EPI_ISL_4978 ···················· ···················· ···················· ···················· ···················· ····················
F 6745 hCoV-19/Hong Kong/HKU-200723-074/2020|EPI_ISL_4978 ···················· ·A·················· ···················· ···················· ···················· ··A·················
F 6746 hCoV-19/Hong Kong/HKU-200723-075/2020|EPI_ISL_4978 ···················· ···················· ···················· ···················· ···················· ····················
F 6747 hCoV-19/Hong Kong/HKU-200723-077/2020|EPI_ISL_4978 ···················· ···················· ···················· ···················· ···················· ····················
F 6748 hCoV-19/Hong Kong/HKU-200723-078/2020|EPI_ISL_4978 ···················· ···················· ···················· ···················· ···················· ····················
F 6749 hCoV-19/Hong Kong/HKU-200723-079/2020|EPI_ISL_4978 ···················· ···················· ···················· ···················· ···················· ····················
F 6750 hCoV-19/Hong Kong/HKU-200723-080/2020|EPI_ISL_4978 ···················· ···················· ···················· ···················· ···················· ····················
F 6751 hCoV-19/Hong Kong/HKU-200723-081/2020|EPI_ISL_4978 ···················· ···················· ···················· ···················· ···················· ····················
F 6752 hCoV-19/Hong Kong/HKU-200723-082/2020|EPI_ISL_4978 ···················· ···················· ···················· ···················· ···················· ····················
F 6753 hCoV-19/Hong Kong/HKU-200723-083/2020|EPI_ISL_4978 ···················· ···················· ···················· ···················· ···················· ····················
F 6754 hCoV-19/Hong Kong/HKU-200723-084/2020|EPI_ISL_4978 ···················· ···················· ···················· ···················· ···················· ····················
F 6755 hCoV-19/Hong Kong/HKU-200723-086/2020|EPI_ISL_4978 ···················· ···················· ···················· ···················· ···················· ····················
F 6756 hCoV-19/Hong Kong/HKU-200723-087/2020|EPI_ISL_4978 ···················· ···················· ···················· ···················· ···················· ····················
F 6757 hCoV-19/Hong Kong/HKU-200723-088/2020|EPI_ISL_4978 ···················· ···················· ···················· ···················· ···················· ····················
F 6758 hCoV-19/Hong Kong/HKU-200723-089/2020|EPI_ISL_4978 ···················· ···················· ···················· ···················· ···················· ····················
F 6759 hCoV-19/Hong Kong/HKU-200723-090/2020|EPI_ISL_4978 ···················· ···················· ···················· ···················· ···················· ····················
F 6760 hCoV-19/Hong Kong/HKU-200723-093/2020|EPI_ISL_4978 ···················· ···················· ···················· ···················· ···················· ····················
F 6761 hCoV-19/Hong Kong/HKU-200723-094/2020|EPI_ISL_4978 ···················· ···················· ···················· ···················· ···················· ····················
F 6762 hCoV-19/Hong Kong/HKU-200723-095/2020|EPI_ISL_4978 ···················· ···················· ···················· ···················· ···················· ····················
F 6763 hCoV-19/Hong Kong/HKU-200723-096/2020|EPI_ISL_4978 ···················· ···················· ···················· ···················· ···················· ····················
F 6764 hCoV-19/Hong Kong/HKU-200723-097/2020|EPI_ISL_4978 ···················· ···················· ···················· ···················· ···················· ····················
F 6765 hCoV-19/Hong Kong/HKU-200723-098/2020|EPI_ISL_4978 ···················· ···················· ···················· ···················· ···················· ····················
F 6766 hCoV-19/Hong Kong/HKU-200723-099/2020|EPI_ISL_4978 ···················· ···················· ···················· ···················· ···················· ····················
F 6767 hCoV-19/Hong Kong/HKU-200723-100/2020|EPI_ISL_4978 ···················· ···················· ···················· ···················· ···················· ····················
F 6768 hCoV-19/Hong Kong/HKU-200723-101/2020|EPI_ISL_4978 ···················· ···················· ···················· ···················· ···················· ····················
F 6769 hCoV-19/Hong Kong/HKU-200723-102/2020|EPI_ISL_4978 ···················· ···················· ···················· ···················· ···················· ····················
F 6770 hCoV-19/Hong Kong/HKU-200723-103/2020|EPI_ISL_4978 ···················· ···················· ···················· ···················· ···················· ····················
F 6782 hCoV-19/USA/MI-MDHHS-SC21094/2020|EPI_ISL_507870|2 ···················· ···················· ···················· ···················· ···················· ····················
F 6783 hCoV-19/USA/MI-MDHHS-SC21095/2020|EPI_ISL_507871|2 ···················· ···················· ···················· ···················· ···················· ····················
F 6784 hCoV-19/USA/MI-MDHHS-SC21098/2020|EPI_ISL_507874|2 ···················· ···················· ···················· ···················· ···················· ····················
F 6785 hCoV-19/USA/MI-MDHHS-SC21103/2020|EPI_ISL_507879|2 ···················· ···················· ···················· ···················· ···················· ····················
F 6786 hCoV-19/USA/MI-MDHHS-SC21105/2020|EPI_ISL_507881|2 ···················· ···················· ···················· ···················· ···················· ····················
F 6787 hCoV-19/USA/MI-MDHHS-SC21106/2020|EPI_ISL_507882|2 ···················· ···················· ···················· ···················· ···················· ····················
F 6788 hCoV-19/USA/MI-MDHHS-SC21107/2020|EPI_ISL_507883|2 ···················· ···················· ···················· ···················· ···················· ····················
F 6789 hCoV-19/USA/MI-MDHHS-SC21108/2020|EPI_ISL_507884|2 ···················· ···················· ···················· ···················· ···················· ····················
F 6790 hCoV-19/USA/MI-MDHHS-SC21109/2020|EPI_ISL_507885|2 ···················· ···················· ···················· ···················· ···················· ····················
F 6791 hCoV-19/USA/MI-MDHHS-SC21110/2020|EPI_ISL_507886|2 ···················· ···················· ···················· ···················· ···················· ····················
F 6792 hCoV-19/USA/MI-MDHHS-SC21111/2020|EPI_ISL_507887|2 ···················· ···················· ···················· ···················· ···················· ····················
F 6793 hCoV-19/USA/MI-MDHHS-SC21112/2020|EPI_ISL_507888|2 ···················· ···················· ···················· ···················· ···················· ····················
F 6794 hCoV-19/USA/MI-MDHHS-SC21113/2020|EPI_ISL_507889|2 ···················· ···················· ···················· ···················· ···················· ····················
F 6795 hCoV-19/USA/MI-MDHHS-SC21114/2020|EPI_ISL_507890|2 ···················· ···················· ···················· ···················· ···················· ····················
F 6796 hCoV-19/USA/MI-MDHHS-SC21115/2020|EPI_ISL_507891|2 ···················· ···················· ···················· ···················· ···················· ····················
F 6797 hCoV-19/USA/MI-MDHHS-SC21116/2020|EPI_ISL_507892|2 ···················· ···················· ···················· ···················· ···················· ····················
F 6798 hCoV-19/USA/MI-MDHHS-SC21117/2020|EPI_ISL_507893|2 ···················· ···················· ···················· ···················· ···················· ····················
F 6799 hCoV-19/USA/MI-MDHHS-SC21118/2020|EPI_ISL_507894|2 ···················· ···················· ···················· ···················· ···················· ····················
F 6800 hCoV-19/USA/MI-MDHHS-SC21119/2020|EPI_ISL_507895|2 ···················· ···················· ···················· ···················· ···················· ····················
F 6801 hCoV-19/USA/MI-MDHHS-SC21120/2020|EPI_ISL_507896|2 ···················· ···················· ···················· ···················· ···················· ····················
F 6802 hCoV-19/USA/MI-MDHHS-SC21121/2020|EPI_ISL_507897|2 ···················· ···················· ···················· ···················· ···················· ····················

| | | | | | | | | |
|---|---|---|---|---|---|---|---|---|
| F | 6803 | hCoV-19/USA/MI-MDHHS-SC21122/2020\|EPI_ISL_507898\|2 | .................... | .................... | .................... | .................... | .................... | .................... |
| F | 6804 | hCoV-19/USA/MI-MDHHS-SC21123/2020\|EPI_ISL_507899\|2 | .................... | .................... | .................... | .................... | .................... | .................... |
| F | 6805 | hCoV-19/Portugal/PT0741/2020\|EPI_ISL_511000\|2020-0 | .................... | .................... | .................... | .................... | .T.................. | .................... |
| F | 6806 | hCoV-19/Portugal/PT0742/2020\|EPI_ISL_511001\|2020-0 | .................... | .................... | .................... | .................... | .................... | .................... |
| F | 6807 | hCoV-19/Portugal/PT0744/2020\|EPI_ISL_511003\|2020-0 | .................... | .................... | .................... | .................... | .................... | .................... |
| F | 6808 | hCoV-19/Portugal/PT0745/2020\|EPI_ISL_511004\|2020-0 | .................... | .................... | .................... | .................... | .................... | .................... |
| F | 6809 | hCoV-19/Portugal/PT0747a/2020\|EPI_ISL_511006\|2020- | .................... | .................... | .................... | .................... | .................... | .................... |
| F | 6810 | hCoV-19/Portugal/PT0801/2020\|EPI_ISL_511060\|2020-0 | .................... | .................... | .................... | .................... | .................... | .................... |
| F | 6811 | hCoV-19/Portugal/PT0802/2020\|EPI_ISL_511061\|2020-0 | .................... | .................... | .................... | .................... | .................... | .................... |
| F | 6812 | hCoV-19/Portugal/PT0803/2020\|EPI_ISL_511062\|2020-0 | .................... | .................... | .................... | .................... | .................... | .................... |
| F | 6813 | hCoV-19/Portugal/PT0804/2020\|EPI_ISL_511063\|2020-0 | .................... | .................... | .................... | .................... | .................... | .................... |
| F | 6814 | hCoV-19/Portugal/PT0805/2020\|EPI_ISL_511064\|2020-0 | .................... | .................... | .................... | .................... | .................... | .................... |
| F | 6815 | hCoV-19/Portugal/PT0806/2020\|EPI_ISL_511065\|2020-0 | .................... | .................... | .................... | .................... | .................... | .................... |
| F | 6816 | hCoV-19/Portugal/PT0807/2020\|EPI_ISL_511066\|2020-0 | .................... | .................... | .................... | .................... | .................... | .................... |
| F | 6817 | hCoV-19/Portugal/PT0808/2020\|EPI_ISL_511067\|2020-0 | .................... | .................... | .................... | .................... | .................... | .................... |
| F | 6818 | hCoV-19/Portugal/PT0809/2020\|EPI_ISL_511068\|2020-0 | .................... | .................... | .................... | .................... | .................... | .................... |
| F | 6819 | hCoV-19/Portugal/PT0810/2020\|EPI_ISL_511069\|2020-0 | .................... | .................... | .................... | .................... | .................... | .................... |
| F | 6820 | hCoV-19/Portugal/PT0811/2020\|EPI_ISL_511070\|2020-0 | .................... | .................... | .................... | .................... | .................... | .................... |
| F | 6821 | hCoV-19/Portugal/PT0812/2020\|EPI_ISL_511071\|2020-0 | .................... | .................... | .................... | .................... | .................... | .................... |
| F | 6822 | hCoV-19/Portugal/PT0813/2020\|EPI_ISL_511072\|2020-0 | .................... | .................... | .................... | .................... | .................... | .................... |
| F | 6823 | hCoV-19/Portugal/PT0814/2020\|EPI_ISL_511073\|2020-0 | .................... | .................... | .................... | .................... | .................... | .................... |
| F | 6824 | hCoV-19/Portugal/PT0815/2020\|EPI_ISL_511074\|2020-0 | .................... | .................... | .................... | .................... | .................... | .................... |
| F | 6825 | hCoV-19/Portugal/PT0817/2020\|EPI_ISL_511076\|2020-0 | .................... | .................... | .................... | .................... | .................... | .................... |
| F | 6826 | hCoV-19/Portugal/PT0818/2020\|EPI_ISL_511077\|2020-0 | .................... | .................... | .................... | .................... | .................... | .................... |
| F | 6827 | hCoV-19/Portugal/PT0820/2020\|EPI_ISL_511079\|2020-0 | .................... | .................... | .................... | .................... | .................... | .................... |
| F | 6828 | hCoV-19/Portugal/PT0821/2020\|EPI_ISL_511080\|2020-0 | .................... | .................... | .................... | .................... | .................... | .................... |
| F | 6829 | hCoV-19/Portugal/PT0823/2020\|EPI_ISL_511082\|2020-0 | .................... | .................... | .................... | .................... | .................... | .................... |
| F | 6830 | hCoV-19/Portugal/PT0824/2020\|EPI_ISL_511083\|2020-0 | .................... | .................... | .................... | .................... | .................... | .................... |
| F | 6831 | hCoV-19/Portugal/PT0826/2020\|EPI_ISL_511085\|2020-0 | .................... | .................... | .................... | .................... | .................... | .................... |
| F | 6832 | hCoV-19/Portugal/PT0827/2020\|EPI_ISL_511086\|2020-0 | .................... | .................... | .................... | .................... | .................... | .................... |
| F | 6833 | hCoV-19/Portugal/PT0828/2020\|EPI_ISL_511087\|2020-0 | .................... | .................... | .................... | .................... | .................... | .................... |
| F | 6834 | hCoV-19/Portugal/PT0830/2020\|EPI_ISL_511089\|2020-0 | .................... | .................... | .................... | .................... | .................... | .................... |
| F | 6835 | hCoV-19/Portugal/PT0831/2020\|EPI_ISL_511090\|2020-0 | .................... | .................... | .................... | .................... | .................... | .................... |
| F | 6836 | hCoV-19/Portugal/PT0832/2020\|EPI_ISL_511091\|2020-0 | .................... | .................... | .................... | .................... | .................... | .................... |
| F | 6837 | hCoV-19/Portugal/PT0833/2020\|EPI_ISL_511092\|2020-0 | .................... | .................... | .................... | .................... | .................... | .................... |
| F | 6838 | hCoV-19/Portugal/PT0834/2020\|EPI_ISL_511093\|2020-0 | .................... | .................... | .................... | .................... | .................... | .................... |
| F | 6839 | hCoV-19/Portugal/PT0835/2020\|EPI_ISL_511094\|2020-0 | .................... | .................... | .................... | .................... | .................... | .................... |
| F | 6840 | hCoV-19/Portugal/PT0836/2020\|EPI_ISL_511095\|2020-0 | .................... | .................... | .................... | .................... | .................... | .................... |
| F | 6841 | hCoV-19/Portugal/PT0837/2020\|EPI_ISL_511096\|2020-0 | .................... | .................... | .................... | .................... | .................... | .................... |
| F | 6842 | hCoV-19/Portugal/PT0838/2020\|EPI_ISL_511097\|2020-0 | .................... | .................... | .................... | .................... | .................... | .................... |
| F | 6843 | hCoV-19/Portugal/PT0839/2020\|EPI_ISL_511098\|2020-0 | .................... | .................... | .................... | .................... | .................... | .................... |
| F | 6844 | hCoV-19/Portugal/PT0840/2020\|EPI_ISL_511099\|2020-0 | .................... | .................... | .................... | .................... | .................... | .................... |
| F | 6845 | hCoV-19/Latvia/088/2020\|EPI_ISL_512313\|2020-06-17 | .................... | .................... | .................... | .................... | .................... | .................... |
| F | 6846 | hCoV-19/Latvia/089/2020\|EPI_ISL_512314\|2020-07-09 | .................... | .................... | .................... | .................... | .................... | .................... |
| F | 6847 | hCoV-19/England/CAMB-1B586B/2020\|EPI_ISL_512332\|20 | .................... | .................... | .................... | .................... | .................... | .................... |
| F | 6848 | hCoV-19/England/CAMB-1B5889/2020\|EPI_ISL_512333\|20 | .................... | .................... | .................... | .................... | .................... | .................... |
| F | 6849 | hCoV-19/England/PORT-2D14CE/2020\|EPI_ISL_512395\|20 | .................... | .................... | .................... | .................... | .................... | .................... |
| F | 6850 | hCoV-19/USA/ME-HETL-J0095/2020\|EPI_ISL_513504\|2020 | .................... | .................... | .................... | .................... | .................... | .................... |
| F | 6851 | hCoV-19/USA/ME-HETL-J0096/2020\|EPI_ISL_513505\|2020 | .................... | .................... | .................... | .................... | .................... | .................... |
| F | 6852 | hCoV-19/USA/ME-HETL-J0097/2020\|EPI_ISL_513506\|2020 | .................... | .................... | .................... | .................... | .................... | .................... |
| F | 6853 | hCoV-19/USA/ME-HETL-J0098/2020\|EPI_ISL_513507\|2020 | .................... | .................... | .................... | .................... | .................... | .................... |
| F | 6854 | hCoV-19/USA/ME-HETL-J0099/2020\|EPI_ISL_513508\|2020 | .................... | .................... | .................... | .................... | .................... | .................... |
| F | 6855 | hCoV-19/USA/ME-HETL-J0100/2020\|EPI_ISL_513509\|2020 | .................... | .................... | .................... | .................... | .................... | .................... |
| F | 6856 | hCoV-19/USA/ME-HETL-J0101/2020\|EPI_ISL_513510\|2020 | .................... | .................... | .................... | .................... | .................... | .................... |
| F | 6857 | hCoV-19/Czech Republic/NRL_2676/2020\|EPI_ISL_51351 | .................... | .................... | .................... | .................... | .................... | .................... |
| F | 6858 | hCoV-19/Czech Republic/NRL_4037/2020\|EPI_ISL_51351 | .................... | .................... | .................... | .................... | .................... | .................... |
| F | 6859 | hCoV-19/Brazil/RJ-INCA-C06/2020\|EPI_ISL_513513\|202 | .................... | .................... | .................... | .................... | .................... | .................... |
| F | 6860 | hCoV-19/Brazil/RJ-INCA-C07/2020\|EPI_ISL_513514\|202 | .................... | .................... | .................... | .................... | .................... | .................... |
| F | 6861 | hCoV-19/USA/WA-UW-4758/2020\|EPI_ISL_513637\|2020-03 | .................... | .................... | .................... | .................... | .................... | .................... |
| F | 6862 | hCoV-19/USA/UT-02759/2020\|EPI_ISL_513645\|2020-04-2 | .................... | .................... | .................... | .................... | .................... | .................... |
| F | 6863 | hCoV-19/USA/UT-02837/2020\|EPI_ISL_513647\|2020-04-2 | .................... | .................... | .................... | .................... | .................... | .................... |
| F | 6864 | hCoV-19/USA/UT-02838/2020\|EPI_ISL_513648\|2020-04-2 | .................... | .................... | .................... | .................... | .................... | .................... |
| F | 6865 | hCoV-19/USA/UT-02842/2020\|EPI_ISL_513652\|2020-04-2 | .................... | .................... | .................... | .................... | .................... | .................... |
| F | 6866 | hCoV-19/USA/UT-02848/2020\|EPI_ISL_513657\|2020-04-2 | .................... | .................... | .................... | .................... | .................... | .................... |
| F | 6867 | hCoV-19/South Korea/KCDC2425/2020\|EPI_ISL_514828\|2 | .................... | .................... | .................... | .................... | .................... | .................... |
| F | 6868 | hCoV-19/South Korea/KCDC2432/2020\|EPI_ISL_514835\|2 | .................... | .................... | .................... | .................... | .................... | .................... |
| F | 6869 | hCoV-19/South Korea/KCDC2433/2020\|EPI_ISL_514836\|2 | .................... | .................... | .................... | .................... | .................... | .................... |
| F | 6870 | hCoV-19/South Korea/KCDC2434/2020\|EPI_ISL_514837\|2 | .................... | .................... | .................... | .................... | .................... | .................... |
| F | 6871 | hCoV-19/South Korea/KCDC2435/2020\|EPI_ISL_514838\|2 | .................... | .................... | .................... | .................... | .................... | .................... |
| F | 6872 | hCoV-19/South Korea/KCDC2436/2020\|EPI_ISL_514839\|2 | .................... | .................... | .................... | .................... | .................... | .................... |
| F | 6873 | hCoV-19/South Korea/KCDC2437/2020\|EPI_ISL_514840\|2 | .................... | .................... | .................... | .................... | .................... | .................... |
| F | 6874 | hCoV-19/South Korea/KCDC2438/2020\|EPI_ISL_514841\|2 | .................... | .................... | .................... | .................... | .................... | .................... |
| F | 6875 | hCoV-19/South Korea/KCDC2439/2020\|EPI_ISL_514842\|2 | .................... | .................... | .................... | .................... | .................... | .................... |
| F | 6876 | hCoV-19/South Korea/KCDC2440/2020\|EPI_ISL_514843\|2 | .................... | .................... | .................... | .................... | .................... | .................... |
| F | 6877 | hCoV-19/South Korea/KCDC2441/2020\|EPI_ISL_514844\|2 | .................... | .................... | .................... | .................... | .................... | .................... |
| F | 6878 | hCoV-19/South Korea/KCDC2442/2020\|EPI_ISL_514845\|2 | .................... | .................... | .................... | .................... | .................... | .................... |
| F | 6879 | hCoV-19/South Korea/KCDC2564/2020\|EPI_ISL_514967\|2 | .................... | .................... | .................... | .................... | .................... | .................... |
| F | 6880 | hCoV-19/South Korea/KCDC2565/2020\|EPI_ISL_514968\|2 | .................... | .................... | .................... | .................... | .................... | .................... |
| F | 6881 | hCoV-19/South Korea/KCDC2566/2020\|EPI_ISL_514969\|2 | .................... | .................... | .................... | .................... | .................... | .................... |
| F | 6882 | hCoV-19/South Korea/KCDC2567/2020\|EPI_ISL_514970\|2 | .................... | .................... | .................... | .................... | .................... | .................... |

| | | | | | | | | |
|---|---|---|---|---|---|---|---|---|
| F | 6883 | hCoV-19/South Korea/KCDC2568/2020\|EPI_ISL_514971\|2 | ···················· | ···················· | ···················· | ···················· | ···················· | ···················· |
| F | 6884 | hCoV-19/South Korea/KCDC2569/2020\|EPI_ISL_514972\|2 | ···················· | ···················· | ···················· | ···················· | ···················· | ···················· |
| F | 6885 | hCoV-19/South Korea/KCDC2570/2020\|EPI_ISL_514973\|2 | ···················· | ···················· | ···················· | ···················· | ···················· | ···················· |
| F | 6886 | hCoV-19/South Korea/KCDC2571/2020\|EPI_ISL_514974\|2 | ···················· | ···················· | ···················· | ···················· | ···················· | ···················· |
| F | 6887 | hCoV-19/South Korea/KCDC2572/2020\|EPI_ISL_514975\|2 | ···················· | ···················· | ···················· | ···················· | ···················· | ···················· |
| F | 6888 | hCoV-19/South Korea/KCDC2573/2020\|EPI_ISL_514976\|2 | ···················· | ···················· | ···················· | ···················· | ···················· | ···················· |
| F | 6889 | hCoV-19/South Korea/KCDC2574/2020\|EPI_ISL_514977\|2 | ···················· | ···················· | ···················· | ···················· | ···················· | ···················· |
| F | 6890 | hCoV-19/South Korea/KCDC2575/2020\|EPI_ISL_514978\|2 | ···················· | ···················· | ···················· | ···················· | ···················· | ···················· |
| F | 6891 | hCoV-19/South Korea/KCDC2576/2020\|EPI_ISL_514979\|2 | ···················· | ···················· | ···················· | ···················· | ···················· | ···················· |
| F | 6892 | hCoV-19/South Korea/KCDC2577/2020\|EPI_ISL_514980\|2 | ···················· | ···················· | ···················· | ···················· | ···················· | ···················· |
| F | 6893 | hCoV-19/South Korea/KCDC2578/2020\|EPI_ISL_514981\|2 | ···················· | ···················· | ···················· | ···················· | ···················· | ···················· |
| F | 6894 | hCoV-19/South Korea/KCDC2579/2020\|EPI_ISL_514982\|2 | ···················· | ···················· | ···················· | ···················· | ···················· | ···················· |
| F | 6895 | hCoV-19/South Korea/KCDC2580/2020\|EPI_ISL_514983\|2 | ···················· | ···················· | ···················· | ···················· | ···················· | ···················· |
| F | 6896 | hCoV-19/South Korea/KCDC2581/2020\|EPI_ISL_514984\|2 | ···················· | ···················· | ···················· | ···················· | ···················· | ···················· |
| F | 6897 | hCoV-19/South Korea/KCDC2582/2020\|EPI_ISL_514985\|2 | ···················· | ···················· | ···················· | ···················· | ···················· | ···················· |
| F | 6898 | hCoV-19/South Korea/KCDC2583/2020\|EPI_ISL_514986\|2 | ···················· | ···················· | ···················· | ···················· | ···················· | ···················· |
| F | 6899 | hCoV-19/South Korea/KCDC2584/2020\|EPI_ISL_514987\|2 | ···················· | ···················· | ···················· | ···················· | ···················· | ···················· |
| F | 6900 | hCoV-19/South Korea/KCDC2585/2020\|EPI_ISL_514988\|2 | ···················· | ···················· | ···················· | ···················· | ···················· | ···················· |
| F | 6901 | hCoV-19/South Korea/KCDC2589/2020\|EPI_ISL_514992\|2 | ···················· | ···················· | ···················· | ···················· | ···················· | ···················· |
| F | 6902 | hCoV-19/South Korea/KCDC2590/2020\|EPI_ISL_514993\|2 | ···················· | ···················· | ···················· | ···················· | ···················· | ···················· |
| F | 6903 | hCoV-19/South Korea/KCDC2591/2020\|EPI_ISL_514994\|2 | ···················· | ···················· | ···················· | ···················· | ···················· | ···················· |
| F | 6904 | hCoV-19/South Korea/KCDC2592/2020\|EPI_ISL_514995\|2 | ···················· | ···················· | ···················· | ···················· | ···················· | ···················· |
| F | 6905 | hCoV-19/South Korea/KCDC2593/2020\|EPI_ISL_514996\|2 | ···················· | ···················· | ···················· | ···················· | ···················· | ···················· |
| F | 6906 | hCoV-19/South Korea/KCDC2594/2020\|EPI_ISL_514997\|2 | ···················· | ···················· | ···················· | ···················· | ···················· | ···················· |
| F | 6907 | hCoV-19/South Korea/KCDC2596/2020\|EPI_ISL_514999\|2 | ···················· | ···················· | ···················· | ···················· | ···················· | ···················· |
| F | 6908 | hCoV-19/mink/Denmark/AD38-Farm2/2020\|EPI_ISL_62634 | ···················· | ···················· | ···················· | ···················· | ···················· | ···················· |
| F | 6909 | hCoV-19/mink/Denmark/AD40-Farm3/2020\|EPI_ISL_62634 | ···················· | ···················· | ···················· | ···················· | ···················· | ···················· |
| F | 6910 | hCoV-19/mink/Denmark/AL3-Farm1/2020\|EPI_ISL_626342 | ···················· | ···················· | ···················· | ···················· | ···················· | ···················· |
| F | 6911 | hCoV-19/mink/Denmark/AD3-Farm1/2020\|EPI_ISL_626343 | ···················· | ···················· | ···················· | ···················· | ···················· | ···················· |
| F | 6912 | hCoV-19/mink/Denmark/AD4-Farm1/2020\|EPI_ISL_626344 | ···················· | ···················· | ···················· | ···················· | ···················· | ···················· |
| F | 6913 | hCoV-19/South Korea/KCDC2071/2020\|EPI_ISL_497972\|2 | ···················· | ···················· | ···················· | ···················· | ···················· | ···················· |
| F | 6914 | hCoV-19/South Korea/KCDC2072/2020\|EPI_ISL_497973\|2 | ···················· | ···················· | ···················· | ···················· | ···················· | ···················· |
| F | 6915 | hCoV-19/South Korea/KCDC2074/2020\|EPI_ISL_497975\|2 | ···················· | ···················· | ···················· | ···················· | ···················· | ···················· |
| F | 6916 | hCoV-19/South Korea/KCDC2078/2020\|EPI_ISL_497979\|2 | ···················· | ···················· | ···················· | ···················· | ···················· | ···················· |
| F | 6918 | hCoV-19/England/BRIS-13107E/2020\|EPI_ISL_501556\|20 | ···················· | ···················· | ···················· | ···················· | ···················· | ···················· |
| F | 6919 | hCoV-19/England/BRIS-131005/2020\|EPI_ISL_501559\|20 | ···················· | ···················· | ···················· | ···················· | ···················· | ···················· |
| F | 6920 | hCoV-19/England/BRIS-130FDC/2020\|EPI_ISL_501560\|20 | ···················· | ···················· | ···················· | ···················· | ···················· | ···················· |
| F | 6921 | hCoV-19/England/BRIS-130E66/2020\|EPI_ISL_501561\|20 | ···················· | ···················· | ···················· | ···················· | ···················· | ···················· |
| F | 6922 | hCoV-19/England/BRIS-130DA5/2020\|EPI_ISL_501563\|20 | ···················· | ···················· | ···················· | ···················· | ···················· | ···················· |
| F | 6923 | hCoV-19/England/BRIS-1310BA/2020\|EPI_ISL_501564\|20 | ···················· | ···················· | ···················· | ···················· | ···················· | ···················· |
| F | 6924 | hCoV-19/England/BRIS-130FEB/2020\|EPI_ISL_501565\|20 | ···················· | ···················· | ···················· | ···················· | ···················· | ···················· |
| F | 6925 | hCoV-19/England/BRIS-130FFA/2020\|EPI_ISL_501566\|20 | ···················· | ···················· | ···················· | ···················· | ···················· | ···················· |
| F | 6926 | hCoV-19/England/BRIS-130F63/2020\|EPI_ISL_501567\|20 | ···················· | ···················· | ···················· | ···················· | ···················· | ···················· |
| F | 6927 | hCoV-19/England/BRIS-130DB4/2020\|EPI_ISL_501568\|20 | ···················· | ···················· | ···················· | ···················· | ···················· | ···················· |
| F | 6928 | hCoV-19/England/BRIS-130E93/2020\|EPI_ISL_501570\|20 | ···················· | ···················· | ···················· | ···················· | ···················· | ···················· |
| F | 6929 | hCoV-19/England/BRIS-130D69/2020\|EPI_ISL_501572\|20 | ···················· | ···················· | ···················· | ···················· | ···················· | ···················· |
| F | 6930 | hCoV-19/England/BRIS-130E0C/2020\|EPI_ISL_501573\|20 | ···················· | ···················· | ···················· | ···················· | ···················· | ···················· |
| F | 6931 | hCoV-19/England/BRIS-130DC3/2020\|EPI_ISL_501574\|20 | ···················· | ···················· | ···················· | ···················· | ···················· | ···················· |
| F | 6932 | hCoV-19/England/BRIS-130D96/2020\|EPI_ISL_501575\|20 | ···················· | ···················· | ···················· | ···················· | ···················· | ···················· |
| F | 6933 | hCoV-19/England/BRIS-130EEE/2020\|EPI_ISL_501578\|20 | ···················· | ···················· | ···················· | ···················· | ···················· | ···················· |
| F | 6934 | hCoV-19/England/BRIS-130E75/2020\|EPI_ISL_501580\|20 | ···················· | ···················· | ···················· | ···················· | ···················· | ···················· |
| F | 6935 | hCoV-19/England/BRIS-131032/2020\|EPI_ISL_501581\|20 | ···················· | ···················· | ···················· | ···················· | ···················· | ···················· |
| F | 6936 | hCoV-19/England/BRIS-130FBE/2020\|EPI_ISL_501583\|20 | ···················· | ···················· | ···················· | ···················· | ···················· | ···················· |
| F | 6937 | hCoV-19/England/BRIS-130F27/2020\|EPI_ISL_501586\|20 | ···················· | ···················· | ···················· | ···················· | ···················· | ···················· |
| F | 6938 | hCoV-19/England/BRIS-130E1B/2020\|EPI_ISL_501588\|20 | ···················· | ···················· | ···················· | ···················· | ···················· | ···················· |
| F | 6939 | hCoV-19/England/BRIS-130EA2/2020\|EPI_ISL_501589\|20 | ···················· | ···················· | ···················· | ···················· | ···················· | ···················· |
| F | 6940 | hCoV-19/England/BRIS-130F90/2020\|EPI_ISL_501590\|20 | ···················· | ···················· | ···················· | ···················· | ···················· | ···················· |
| F | 6941 | hCoV-19/England/BRIS-13106F/2020\|EPI_ISL_501591\|20 | ···················· | ···················· | ···················· | ···················· | ···················· | ···················· |
| F | 6942 | hCoV-19/England/BRIS-130E57/2020\|EPI_ISL_501592\|20 | ···················· | ···················· | ···················· | ···················· | ···················· | ···················· |
| F | 6943 | hCoV-19/England/BRIS-1310D8/2020\|EPI_ISL_501593\|20 | ···················· | ···················· | ···················· | ···················· | ···················· | ···················· |
| F | 6944 | hCoV-19/England/BRIS-130F72/2020\|EPI_ISL_501595\|20 | ···················· | ···················· | ···················· | ···················· | ···················· | ···················· |
| F | 6945 | hCoV-19/England/BRIS-130E48/2020\|EPI_ISL_501596\|20 | ···················· | ···················· | ···················· | ···················· | ···················· | ···················· |
| F | 6946 | hCoV-19/England/BRIS-130EC0/2020\|EPI_ISL_501597\|20 | ···················· | ···················· | ···················· | ···················· | ···················· | ···················· |
| F | 6947 | hCoV-19/England/BRIS-130E2A/2020\|EPI_ISL_501598\|20 | ···················· | ···················· | ···················· | ···················· | ···················· | ···················· |
| F | 6948 | hCoV-19/England/BRIS-130DE1/2020\|EPI_ISL_501599\|20 | ···················· | ···················· | ···················· | ···················· | ···················· | ···················· |
| F | 6949 | hCoV-19/Brazil/PE-COV0260/2020\|EPI_ISL_502875\|2020 | ···················· | ···················· | ···················· | ···················· | ···················· | ···················· |
| F | 6954 | hCoV-19/USA/CA-ALSR-1831/2020\|EPI_ISL_512200\|2020- | ···················· | ·A·················· | ···················· | ···················· | ···················· | ···················· |
| F | 6955 | hCoV-19/USA/CA-ALSR-1832/2020\|EPI_ISL_512201\|2020- | ···················· | ···················· | ···················· | ···················· | ···················· | ···················· |
| F | 6956 | hCoV-19/USA/CA-ALSR-1837/2020\|EPI_ISL_512203\|2020- | ···················· | ·A·················· | ···················· | ···················· | ···················· | ···················· |
| F | 6957 | hCoV-19/USA/CA-ALSR-1841/2020\|EPI_ISL_512204\|2020- | ···················· | ·A·················· | ···················· | ···················· | ···················· | ···················· |
| F | 6958 | hCoV-19/USA/CA-ALSR-1845/2020\|EPI_ISL_512206\|2020- | ···················· | ···················· | ···················· | ···················· | ···················· | ···················· |
| F | 6959 | hCoV-19/USA/CA-ALSR-1848/2020\|EPI_ISL_512207\|2020- | ···················· | ·A·················· | ···················· | ···················· | ···················· | ···················· |
| F | 6960 | hCoV-19/USA/CA-ALSR-1854/2020\|EPI_ISL_512208\|2020- | ···················· | ···················· | ···················· | ···················· | ···················· | ···················· |
| F | 6961 | hCoV-19/USA/CA-ALSR-1859/2020\|EPI_ISL_512209\|2020- | ···················· | ·A·················· | ···················· | ···················· | ···················· | ···················· |
| F | 6962 | hCoV-19/USA/CA-ALSR-1862/2020\|EPI_ISL_512211\|2020- | ···················· | ·A·················· | ···················· | ···················· | ···················· | ···················· |
| F | 6963 | hCoV-19/USA/CA-ALSR-1866/2020\|EPI_ISL_512212\|2020- | ···················· | ·A·················· | ···················· | ···················· | ···················· | ···················· |
| F | 6964 | hCoV-19/USA/CA-ALSR-1869/2020\|EPI_ISL_512213\|2020- | ···················· | ·A·················· | ···················· | ···················· | ···················· | ···················· |
| F | 6965 | hCoV-19/USA/CA-ALSR-1876/2020\|EPI_ISL_512214\|2020- | ···················· | ···················· | ···················· | ···················· | ···················· | ···················· |
| F | 6966 | hCoV-19/USA/CA-ALSR-1877/2020\|EPI_ISL_512215\|2020- | ···················· | ·A·················· | ···················· | ···················· | ···················· | ···················· |
| F | 6967 | hCoV-19/USA/CA-ALSR-1889/2020\|EPI_ISL_512220\|2020- | ···················· | ·A·················· | ···················· | ···················· | ···················· | ···················· |

| | | | | | | | |
|---|---|---|---|---|---|---|---|
| F | 6968 hCoV-19/USA/CA-ALSR-1894/2020\|EPI_ISL_512221\|2020- | ···················· | ·A·················· | ···················· | ···················· | ···················· | ···················· |
| F | 6969 hCoV-19/USA/CA-ALSR-1898/2020\|EPI_ISL_512222\|2020- | ···················· | ·A·················· | ···················· | ···················· | ···················· | ···················· |
| F | 6970 hCoV-19/USA/CA-ALSR-1899/2020\|EPI_ISL_512223\|2020- | ···················· | ·A·················· | ···················· | ···················· | ···················· | ···················· |
| F | 6971 hCoV-19/USA/CA-ALSR-1900/2020\|EPI_ISL_512224\|2020- | ···················· | ···················· | ···················· | ···················· | ···················· | ···················· |
| F | 6972 hCoV-19/USA/CA-ALSR-1903/2020\|EPI_ISL_512225\|2020- | ···················· | ·A·················· | ···················· | ···················· | ···················· | ···················· |
| F | 6973 hCoV-19/USA/CA-ALSR-1904/2020\|EPI_ISL_512226\|2020- | ···················· | ·A·················· | ···················· | ···················· | ···················· | ···················· |
| F | 6974 hCoV-19/USA/CA-ALSR-1906/2020\|EPI_ISL_512227\|2020- | ···················· | ·A·················· | ···················· | ···················· | ···················· | ···················· |
| F | 6975 hCoV-19/USA/CA-ALSR-1907/2020\|EPI_ISL_512228\|2020- | ···················· | ·A·················· | ···················· | ···················· | ···················· | ···················· |
| F | 6976 hCoV-19/USA/CA-ALSR-1910/2020\|EPI_ISL_512229\|2020- | ···················· | ···················· | ···················· | ···················· | ···················· | ···················· |
| F | 6977 hCoV-19/USA/CA-ALSR-1912/2020\|EPI_ISL_512230\|2020- | ···················· | ·A·················· | ···················· | ···················· | ···················· | ···················· |
| F | 6978 hCoV-19/USA/CA-ALSR-1915/2020\|EPI_ISL_512231\|2020- | ···················· | ·A·················· | ···················· | ···················· | ···················· | ···················· |
| F | 6979 hCoV-19/USA/CA-ALSR-1916/2020\|EPI_ISL_512232\|2020- | ···················· | ···················· | ···················· | ···················· | ···················· | ···················· |
| F | 6980 hCoV-19/USA/CA-ALSR-1917/2020\|EPI_ISL_512233\|2020- | ···················· | ···················· | ···················· | ···················· | ···················· | ···················· |
| F | 6981 hCoV-19/USA/CA-ALSR-1924/2020\|EPI_ISL_512235\|2020- | ···················· | ·A·················· | ···················· | ···················· | ···················· | ···················· |
| F | 6982 hCoV-19/USA/CA-ALSR-1925/2020\|EPI_ISL_512236\|2020- | ···················· | ·A·················· | ···················· | ···················· | ···················· | ···················· |
| F | 6983 hCoV-19/USA/CA-ALSR-1926/2020\|EPI_ISL_512237\|2020- | ···················· | ·A·················· | ···················· | ···················· | ···················· | ···················· |
| F | 6984 hCoV-19/USA/CA-ALSR-1927/2020\|EPI_ISL_512238\|2020- | ···················· | ·A·················· | ···················· | ···················· | ···················· | ···················· |
| F | 6985 hCoV-19/USA/CA-ALSR-1928/2020\|EPI_ISL_512239\|2020- | ···················· | ·A·················· | ···················· | ···················· | ···················· | ···················· |
| F | 6986 hCoV-19/USA/CA-ALSR-1929/2020\|EPI_ISL_512240\|2020- | ···················· | ·A·················· | ···················· | ···················· | ···················· | ···················· |
| F | 6987 hCoV-19/USA/CA-ALSR-1930/2020\|EPI_ISL_512241\|2020- | ···················· | ·A·················· | ···················· | ···················· | ···················· | ···················· |
| F | 6988 hCoV-19/USA/CA-ALSR-1933/2020\|EPI_ISL_512242\|2020- | ···················· | ·A·················· | ···················· | ···················· | ···················· | ···················· |
| F | 6989 hCoV-19/USA/CA-ALSR-1935/2020\|EPI_ISL_512243\|2020- | ···················· | ·A·················· | ···················· | ···················· | ···················· | ···················· |
| F | 6990 hCoV-19/USA/CA-ALSR-1938/2020\|EPI_ISL_512244\|2020- | ···················· | ·A·················· | ···················· | ···················· | ···················· | ···················· |
| F | 6991 hCoV-19/USA/CA-ALSR-1939/2020\|EPI_ISL_512245\|2020- | ···················· | ·A·················· | ···················· | ···················· | ···················· | ···················· |
| F | 6992 hCoV-19/USA/CA-ALSR-1940/2020\|EPI_ISL_512246\|2020- | ···················· | ·A·················· | ···················· | ···················· | ···················· | ···················· |
| F | 6993 hCoV-19/USA/CA-ALSR-1941/2020\|EPI_ISL_512247\|2020- | ···················· | ·A·················· | ···················· | ···················· | ···················· | ···················· |
| F | 6994 hCoV-19/USA/CA-ALSR-1943/2020\|EPI_ISL_512248\|2020- | ···················· | ·A·················· | ···················· | ···················· | ···················· | ···················· |
| F | 6995 hCoV-19/USA/CA-ALSR-1945/2020\|EPI_ISL_512250\|2020- | ···················· | ·A·················· | ···················· | ···················· | ···················· | ···················· |
| F | 6996 hCoV-19/USA/CA-ALSR-1946/2020\|EPI_ISL_512251\|2020- | ···················· | ·A·················· | ···················· | ···················· | ···················· | ···················· |
| F | 6997 hCoV-19/USA/CA-ALSR-1947/2020\|EPI_ISL_512252\|2020- | ···················· | ·A·················· | ···················· | ···················· | ···················· | ···················· |
| F | 6998 hCoV-19/USA/CA-ALSR-1948/2020\|EPI_ISL_512253\|2020- | ···················· | ·A·················· | ···················· | ···················· | ···················· | ···················· |
| F | 6999 hCoV-19/USA/CA-ALSR-1949/2020\|EPI_ISL_512254\|2020- | ···················· | ·A·················· | ···················· | ···················· | ···················· | ···················· |
| F | 7000 hCoV-19/USA/CA-ALSR-1951/2020\|EPI_ISL_512256\|2020- | ···················· | ·A·················· | ···················· | ···················· | ···················· | ···················· |
| F | 7001 hCoV-19/USA/CA-ALSR-1952/2020\|EPI_ISL_512257\|2020- | ···················· | ·A·················· | ···················· | ···················· | ···················· | ···················· |
| F | 7002 hCoV-19/USA/CA-ALSR-1954/2020\|EPI_ISL_512259\|2020- | ···················· | ·A·················· | ···················· | ···················· | ···················· | ···················· |
| F | 7003 hCoV-19/USA/CA-ALSR-1955/2020\|EPI_ISL_512260\|2020- | ···················· | ·A·················· | ···················· | ···················· | ···················· | ···················· |
| F | 7004 hCoV-19/USA/CA-ALSR-1957/2020\|EPI_ISL_512261\|2020- | ···················· | ·A·················· | ···················· | ···················· | ···················· | ···················· |
| F | 7005 hCoV-19/USA/CA-ALSR-1959/2020\|EPI_ISL_512262\|2020- | ···················· | ·A·················· | ···················· | ···················· | ···················· | ···················· |
| F | 7006 hCoV-19/USA/CA-ALSR-1961/2020\|EPI_ISL_512264\|2020- | ···················· | ·A·················· | ···················· | ···················· | ···················· | ···················· |
| F | 7007 hCoV-19/USA/CA-ALSR-1962/2020\|EPI_ISL_512265\|2020- | ···················· | ·A·················· | ···················· | ···················· | ···················· | ···················· |
| F | 7008 hCoV-19/USA/CA-ALSR-1963/2020\|EPI_ISL_512266\|2020- | ···················· | ·A·················· | ···················· | ···················· | ···················· | ···················· |
| F | 7009 hCoV-19/USA/CA-ALSR-1967/2020\|EPI_ISL_512267\|2020- | ···················· | ·A·················· | ···················· | ···················· | ···················· | ···················· |
| F | 7010 hCoV-19/USA/CA-ALSR-1970/2020\|EPI_ISL_512268\|2020- | ···················· | ·A·················· | ···················· | ···················· | ···················· | ···················· |
| F | 7011 hCoV-19/USA/CA-ALSR-1971/2020\|EPI_ISL_512269\|2020- | ···················· | ·A·················· | ···················· | ···················· | ···················· | ···················· |
| F | 7012 hCoV-19/USA/ME-HETL-J0091/2020\|EPI_ISL_513500\|2020 | ···················· | ···················· | ···················· | ···················· | ···················· | ···················· |
| F | 7013 hCoV-19/USA/ME-HETL-J0092/2020\|EPI_ISL_513501\|2020 | ···················· | ···················· | ···················· | ···················· | ···················· | ···················· |
| F | 7014 hCoV-19/USA/ME-HETL-J0093/2020\|EPI_ISL_513502\|2020 | ···················· | ···················· | ···················· | ···················· | ···················· | ···················· |
| F | 7015 hCoV-19/USA/ME-HETL-J0094/2020\|EPI_ISL_513503\|2020 | ···················· | ···················· | ···················· | ···················· | ···················· | ···················· |
| F | 7016 hCoV-19/Brazil/RJ-INCA-C12/2020\|EPI_ISL_513515\|202 | ···················· | ···················· | ···················· | ···················· | ···················· | ···················· |
| F | 7017 hCoV-19/Brazil/RJ-INCA-C15/2020\|EPI_ISL_513518\|202 | ···················· | ···················· | ···················· | ···················· | ···················· | ···················· |
| F | 7018 hCoV-19/Brazil/RJ-INCA-C181/2020\|EPI_ISL_513519\|20 | ···················· | ···················· | ···················· | ···················· | ···················· | ···················· |
| F | 7019 hCoV-19/Brazil/RJ-INCA-C182/2020\|EPI_ISL_513520\|20 | ···················· | ···················· | ···················· | ···················· | ···················· | ···················· |
| F | 7020 hCoV-19/Brazil/RJ-INCA-C183/2020\|EPI_ISL_513521\|20 | ···················· | ···················· | ···················· | ···················· | ···················· | ···················· |
| F | 7022 hCoV-19/Brazil/RJ-INCA-C190/2020\|EPI_ISL_513524\|20 | ···················· | ···················· | ···················· | ···················· | ···················· | ···················· |
| F | 7023 hCoV-19/Brazil/RJ-INCA-C194/2020\|EPI_ISL_513525\|20 | ···················· | ···················· | ···················· | ···················· | ···················· | ···················· |
| F | 7024 hCoV-19/Brazil/RJ-INCA-C197/2020\|EPI_ISL_513526\|20 | ···················· | ···················· | ···················· | ···················· | ···················· | ···················· |
| F | 7025 hCoV-19/Brazil/RJ-INCA-C199/2020\|EPI_ISL_513527\|20 | ···················· | ···················· | ···················· | ···················· | ···················· | ···················· |
| F | 7026 hCoV-19/Brazil/RJ-INCA-C202/2020\|EPI_ISL_513529\|20 | ···················· | ···················· | ···················· | ···················· | ···················· | ···················· |
| F | 7027 hCoV-19/Brazil/RJ-INCA-C22/2020\|EPI_ISL_513530\|202 | ···················· | ···················· | ···················· | ···················· | ···················· | ···················· |
| F | 7028 hCoV-19/Brazil/RJ-INCA-C23/2020\|EPI_ISL_513531\|202 | ···················· | ···················· | ···················· | ···················· | ···················· | ···················· |
| F | 7029 hCoV-19/Brazil/RJ-INCA-C24/2020\|EPI_ISL_513532\|202 | ···················· | ···················· | ···················· | ···················· | ···················· | ···················· |
| F | 7030 hCoV-19/Brazil/RJ-INCA-C25/2020\|EPI_ISL_513533\|202 | ···················· | ···················· | ···················· | ···················· | ···················· | ···················· |
| F | 7031 hCoV-19/Brazil/RJ-INCA-C26/2020\|EPI_ISL_513534\|202 | ···················· | ···················· | ···················· | ···················· | ···················· | ···················· |
| F | 7032 hCoV-19/Brazil/RJ-INCA-C27/2020\|EPI_ISL_513535\|202 | ···················· | ···················· | ···················· | ···················· | ···················· | ···················· |
| F | 7033 hCoV-19/Brazil/RJ-INCA-C28/2020\|EPI_ISL_513536\|202 | ···················· | ···················· | ···················· | ···················· | ···················· | ···················· |
| F | 7034 hCoV-19/Brazil/RJ-INCA-C29/2020\|EPI_ISL_513537\|202 | ···················· | ···················· | ···················· | ···················· | ···················· | ···················· |
| F | 7035 hCoV-19/Brazil/RJ-INCA-C30/2020\|EPI_ISL_513538\|202 | ···················· | ···················· | ···················· | ···················· | ···················· | ···················· |
| F | 7036 hCoV-19/Brazil/RJ-INCA-C31/2020\|EPI_ISL_513539\|202 | ···················· | ···················· | ···················· | ···················· | ···················· | ···················· |
| F | 7037 hCoV-19/Brazil/RJ-INCA-C32/2020\|EPI_ISL_513540\|202 | ···················· | ···················· | ···················· | ···················· | ···················· | ···················· |
| F | 7038 hCoV-19/Brazil/RJ-INCA-C33/2020\|EPI_ISL_513541\|202 | ···················· | ···················· | ···················· | ···················· | ···················· | ···················· |
| F | 7039 hCoV-19/Brazil/RJ-INCA-C34/2020\|EPI_ISL_513542\|202 | ···················· | ···················· | ···················· | ···················· | ···················· | ···················· |
| F | 7040 hCoV-19/Brazil/RJ-INCA-C35/2020\|EPI_ISL_513543\|202 | ···················· | ···················· | ···················· | ···················· | ···················· | ···················· |
| F | 7041 hCoV-19/Brazil/RJ-INCA-C36/2020\|EPI_ISL_513544\|202 | ···················· | ···················· | ···················· | ···················· | ···················· | ···················· |
| F | 7042 hCoV-19/Brazil/RJ-INCA-C37/2020\|EPI_ISL_513545\|202 | ···················· | ···················· | ···················· | ···················· | ···················· | ···················· |
| F | 7043 hCoV-19/Brazil/RJ-INCA-C38/2020\|EPI_ISL_513546\|202 | ···················· | ···················· | ···················· | ···················· | ···················· | ···················· |
| F | 7044 hCoV-19/Brazil/RJ-INCA-C39/2020\|EPI_ISL_513547\|202 | ···················· | ···················· | ···················· | ···················· | ···················· | ···················· |
| F | 7045 hCoV-19/Brazil/RJ-INCA-C40/2020\|EPI_ISL_513548\|202 | ···················· | ···················· | ···················· | ···················· | ···················· | ···················· |
| F | 7046 hCoV-19/Brazil/RJ-INCA-C41/2020\|EPI_ISL_513549\|202 | ···················· | ···················· | ···················· | ···················· | ···················· | ···················· |
| F | 7047 hCoV-19/Brazil/RJ-INCA-C42/2020\|EPI_ISL_513550\|202 | ···················· | ···················· | ···················· | ···················· | ···················· | ···················· |
| F | 7048 hCoV-19/Brazil/RJ-INCA-C44/2020\|EPI_ISL_513551\|202 | ···················· | ···················· | ···················· | ···················· | ···················· | ···················· |

| | | | | | | | | |
|---|---|---|---|---|---|---|---|---|
| F | 7049 | hCoV-19/Brazil/RJ-INCA-C47/2020\|EPI_ISL_513553\|202 | .................... | .................... | .................... | .................... | .................... | .................... |
| F | 7050 | hCoV-19/Brazil/RJ-INCA-C63/2020\|EPI_ISL_513556\|202 | .................... | .................... | .................... | .................... | .................... | .................... |
| F | 7051 | hCoV-19/Brazil/RJ-INCA-C64/2020\|EPI_ISL_513557\|202 | .................... | .................... | .................... | .................... | .................... | .................... |
| F | 7052 | hCoV-19/Brazil/RJ-INCA-C65/2020\|EPI_ISL_513558\|202 | .................... | .................... | .................... | .................... | .................... | .................... |
| F | 7053 | hCoV-19/Brazil/RJ-INCA-C67/2020\|EPI_ISL_513559\|202 | .................... | .................... | .................... | .................... | .................... | .................... |
| F | 7054 | hCoV-19/Brazil/RJ-INCA-C68/2020\|EPI_ISL_513560\|202 | .................... | .................... | .................... | .................... | .................... | .................... |
| F | 7055 | hCoV-19/Brazil/RJ-INCA-C69/2020\|EPI_ISL_513561\|202 | .................... | .................... | .................... | .................... | .................... | .................... |
| F | 7056 | hCoV-19/Brazil/RJ-INCA-C70/2020\|EPI_ISL_513562\|202 | .................... | .................... | .................... | .................... | .................... | .................... |
| F | 7057 | hCoV-19/Brazil/RJ-INCA-C72/2020\|EPI_ISL_513563\|202 | .................... | .................... | .................... | .................... | .................... | .................... |
| F | 7058 | hCoV-19/Brazil/RJ-INCA-C73/2020\|EPI_ISL_513564\|202 | .................... | .................... | .................... | .................... | .................... | .................... |
| F | 7059 | hCoV-19/Brazil/RJ-INCA-C77/2020\|EPI_ISL_513567\|202 | .................... | .................... | .................... | .................... | .................... | .................... |
| F | 7060 | hCoV-19/Brazil/RJ-INCA-C78/2020\|EPI_ISL_513568\|202 | .................... | .................... | .................... | .................... | .................... | .................... |
| F | 7061 | hCoV-19/Brazil/RJ-INCA-C94/2020\|EPI_ISL_513571\|202 | .................... | .................... | .................... | .................... | .................... | .................... |
| F | 7062 | hCoV-19/Brazil/RJ-INCA-C95/2020\|EPI_ISL_513572\|202 | .................... | .................... | .................... | .................... | .................... | .................... |
| F | 7063 | hCoV-19/Brazil/RJ-INCA-I29/2020\|EPI_ISL_513573\|202 | .................... | .................... | .................... | .................... | .................... | .................... |
| F | 7064 | hCoV-19/Brazil/RJ-INCA-I30/2020\|EPI_ISL_513574\|202 | .................... | .................... | .................... | .................... | .................... | .................... |
| F | 7065 | hCoV-19/Brazil/RJ-INCA-I34/2020\|EPI_ISL_513575\|202 | .................... | .................... | .................... | .................... | .................... | .................... |
| F | 7066 | hCoV-19/Brazil/RJ-INCA-I35/2020\|EPI_ISL_513576\|202 | .................... | .................... | .................... | .................... | .................... | .................... |
| F | 7067 | hCoV-19/Brazil/RJ-INCA-I37/2020\|EPI_ISL_513577\|202 | .................... | .................... | .................... | .................... | .................... | .................... |
| F | 7068 | hCoV-19/Brazil/RJ-INCA-I39/2020\|EPI_ISL_513578\|202 | .................... | .................... | .................... | .................... | .................... | .................... |
| F | 7069 | hCoV-19/Brazil/RJ-INCA-I43/2020\|EPI_ISL_513579\|202 | .................... | .................... | .................... | .................... | .................... | .................... |
| F | 7070 | hCoV-19/Brazil/RJ-INCA-I44/2020\|EPI_ISL_513580\|202 | .................... | .................... | .................... | .................... | .................... | .................... |
| F | 7071 | hCoV-19/Brazil/RJ-INCA-I51/2020\|EPI_ISL_513581\|202 | .................... | .................... | .................... | .................... | .................... | .................... |
| F | 7072 | hCoV-19/Brazil/RJ-INCA-I54/2020\|EPI_ISL_513582\|202 | .................... | .................... | .................... | .................... | .................... | .................... |
| F | 7073 | hCoV-19/Brazil/RJ-INCA-I57/2020\|EPI_ISL_513583\|202 | .................... | .................... | .................... | .................... | .................... | .................... |
| F | 7074 | hCoV-19/England/BRIS-133610/2020\|EPI_ISL_513584\|20 | .................... | .................... | .................... | .................... | .................... | .................... |
| F | 7075 | hCoV-19/England/KGHT-2684527/2020\|EPI_ISL_513585\|2 | .................... | .................... | .................... | .................... | .................... | .................... |
| F | 7076 | hCoV-19/England/KGHT-268426C/2020\|EPI_ISL_513587\|2 | .................... | .................... | .................... | .................... | .................... | .................... |
| F | 7077 | hCoV-19/England/KGHT-26844FD/2020\|EPI_ISL_513588\|2 | .................... | .................... | .................... | .................... | .................... | .................... |
| F | 7078 | hCoV-19/DRC/7042/2020\|EPI_ISL_513592\|2020-05-12 | .................... | .................... | .................... | .................... | .................... | .................... |
| F | 7079 | hCoV-19/DRC/4524/2020\|EPI_ISL_513597\|2020-04-30 | .................... | .................... | .................... | .................... | .................... | .................... |
| F | 7080 | hCoV-19/DRC/4541/2020\|EPI_ISL_513598\|2020-04-30 | .................... | .................... | .................... | .................... | .................... | .................... |
| F | 7081 | hCoV-19/South Korea/KCDC2397/2020\|EPI_ISL_514800\|2 | .................... | .................... | .................... | .................... | .................... | .................... |
| F | 7082 | hCoV-19/South Korea/KCDC2398/2020\|EPI_ISL_514801\|2 | .................... | .................... | .................... | .................... | .................... | .................... |
| F | 7083 | hCoV-19/South Korea/KCDC2399/2020\|EPI_ISL_514802\|2 | .................... | .................... | .................... | .................... | .................... | .................... |
| F | 7084 | hCoV-19/South Korea/KCDC2400/2020\|EPI_ISL_514803\|2 | .................... | .................... | .................... | .................... | .................... | .................... |
| F | 7085 | hCoV-19/South Korea/KCDC2401/2020\|EPI_ISL_514804\|2 | .................... | .................... | .................... | .................... | .................... | .................... |
| F | 7086 | hCoV-19/South Korea/KCDC2402/2020\|EPI_ISL_514805\|2 | .................... | .................... | .................... | .................... | .................... | .................... |
| F | 7087 | hCoV-19/South Korea/KCDC2403/2020\|EPI_ISL_514806\|2 | .................... | .................... | .................... | .................... | .................... | .................... |
| F | 7088 | hCoV-19/South Korea/KCDC2404/2020\|EPI_ISL_514807\|2 | .................... | .................... | .................... | .................... | .................... | .................... |
| F | 7089 | hCoV-19/South Korea/KCDC2405/2020\|EPI_ISL_514808\|2 | .................... | .................... | .................... | .................... | .................... | .................... |
| F | 7090 | hCoV-19/South Korea/KCDC2406/2020\|EPI_ISL_514809\|2 | .................... | .................... | .................... | .................... | .................... | .................... |
| F | 7091 | hCoV-19/South Korea/KCDC2407/2020\|EPI_ISL_514810\|2 | .................... | .................... | .................... | .................... | .................... | .................... |
| F | 7092 | hCoV-19/South Korea/KCDC2408/2020\|EPI_ISL_514811\|2 | .................... | .................... | .................... | .................... | .................... | .................... |
| F | 7093 | hCoV-19/South Korea/KCDC2409/2020\|EPI_ISL_514812\|2 | .................... | .................... | .................... | .................... | .................... | .................... |
| F | 7094 | hCoV-19/South Korea/KCDC2410/2020\|EPI_ISL_514813\|2 | .................... | .................... | .................... | .................... | .................... | .................... |
| F | 7095 | hCoV-19/South Korea/KCDC2411/2020\|EPI_ISL_514814\|2 | .................... | .................... | .................... | .................... | .................... | .................... |
| F | 7096 | hCoV-19/South Korea/KCDC2412/2020\|EPI_ISL_514815\|2 | .................... | .................... | .................... | .................... | .................... | .................... |
| F | 7097 | hCoV-19/South Korea/KCDC2413/2020\|EPI_ISL_514816\|2 | .................... | .................... | .................... | .................... | .................... | .................... |
| F | 7098 | hCoV-19/South Korea/KCDC2414/2020\|EPI_ISL_514817\|2 | .................... | .................... | .................... | .................... | .................... | .................... |
| F | 7099 | hCoV-19/South Korea/KCDC2415/2020\|EPI_ISL_514818\|2 | .................... | .................... | .................... | .................... | .................... | .................... |
| F | 7100 | hCoV-19/South Korea/KCDC2416/2020\|EPI_ISL_514819\|2 | .................... | .................... | .................... | .................... | .................... | .................... |
| F | 7101 | hCoV-19/South Korea/KCDC2417/2020\|EPI_ISL_514820\|2 | .................... | .................... | .................... | .................... | .................... | .................... |
| F | 7102 | hCoV-19/South Korea/KCDC2418/2020\|EPI_ISL_514821\|2 | .................... | .................... | .................... | .................... | .................... | .................... |
| F | 7103 | hCoV-19/South Korea/KCDC2419/2020\|EPI_ISL_514822\|2 | .................... | .................... | .................... | .................... | .................... | .................... |
| F | 7104 | hCoV-19/South Korea/KCDC2420/2020\|EPI_ISL_514823\|2 | .................... | .................... | .................... | .................... | .................... | .................... |
| F | 7105 | hCoV-19/South Korea/KCDC2421/2020\|EPI_ISL_514824\|2 | .................... | .................... | .................... | .................... | .................... | .................... |
| F | 7106 | hCoV-19/South Korea/KCDC2422/2020\|EPI_ISL_514825\|2 | .................... | .................... | .................... | .................... | .................... | .................... |
| F | 7107 | hCoV-19/South Korea/KCDC2423/2020\|EPI_ISL_514826\|2 | .................... | .................... | .................... | .................... | .................... | .................... |
| F | 7108 | hCoV-19/South Korea/KCDC2424/2020\|EPI_ISL_514827\|2 | .................... | .................... | .................... | .................... | .................... | .................... |
| F | 7109 | hCoV-19/South Korea/KCDC2426/2020\|EPI_ISL_514829\|2 | .................... | .................... | .................... | .................... | .................... | .................... |
| F | 7110 | hCoV-19/South Korea/KCDC2427/2020\|EPI_ISL_514830\|2 | .................... | .................... | .................... | .................... | .................... | .................... |
| F | 7111 | hCoV-19/South Korea/KCDC2428/2020\|EPI_ISL_514831\|2 | .................... | .................... | .................... | .................... | .................... | .................... |
| F | 7112 | hCoV-19/South Korea/KCDC2429/2020\|EPI_ISL_514832\|2 | .................... | .................... | .................... | .................... | .................... | .................... |
| F | 7113 | hCoV-19/South Korea/KCDC2430/2020\|EPI_ISL_514833\|2 | .................... | .................... | .................... | .................... | .................... | .................... |
| F | 7114 | hCoV-19/South Korea/KCDC2431/2020\|EPI_ISL_514834\|2 | .................... | .................... | .................... | .................... | .................... | .................... |
| F | 7115 | hCoV-19/South Korea/KCDC2443/2020\|EPI_ISL_514846\|2 | .................... | .................... | .................... | .................... | .................... | .................... |
| F | 7116 | hCoV-19/South Korea/KCDC2444/2020\|EPI_ISL_514847\|2 | .................... | .................... | .................... | .................... | .................... | .................... |
| F | 7117 | hCoV-19/South Korea/KCDC2445/2020\|EPI_ISL_514848\|2 | .................... | .................... | .................... | .................... | .................... | .................... |
| F | 7118 | hCoV-19/South Korea/KCDC2446/2020\|EPI_ISL_514849\|2 | .................... | .................... | .................... | .................... | .................... | .................... |
| F | 7119 | hCoV-19/South Korea/KCDC2447/2020\|EPI_ISL_514850\|2 | .................... | .................... | .................... | .................... | .................... | .................... |
| F | 7120 | hCoV-19/South Korea/KCDC2448/2020\|EPI_ISL_514851\|2 | .................... | .................... | .................... | .................... | .................... | .................... |
| F | 7121 | hCoV-19/South Korea/KCDC2449/2020\|EPI_ISL_514852\|2 | .................... | .................... | .................... | .................... | .................... | .................... |
| F | 7122 | hCoV-19/South Korea/KCDC2450/2020\|EPI_ISL_514853\|2 | .................... | .................... | .................... | .................... | .................... | .................... |
| F | 7123 | hCoV-19/South Korea/KCDC2451/2020\|EPI_ISL_514854\|2 | .................... | .................... | .................... | .................... | .................... | .................... |
| F | 7124 | hCoV-19/South Korea/KCDC2452/2020\|EPI_ISL_514855\|2 | .................... | .................... | .................... | .................... | .................... | .................... |
| F | 7125 | hCoV-19/South Korea/KCDC2453/2020\|EPI_ISL_514856\|2 | .................... | .................... | .................... | .................... | .................... | .................... |
| F | 7126 | hCoV-19/South Korea/KCDC2454/2020\|EPI_ISL_514857\|2 | .................... | .................... | .................... | .................... | .................... | .................... |
| F | 7127 | hCoV-19/South Korea/KCDC2455/2020\|EPI_ISL_514858\|2 | .................... | .................... | .................... | .................... | .................... | .................... |
| F | 7128 | hCoV-19/South Korea/KCDC2456/2020\|EPI_ISL_514859\|2 | .................... | .................... | .................... | .................... | .................... | .................... |

F 7129 hCoV-19/South Korea/KCDC2457/2020|EPI_ISL_514860|2 .................... .................... .................... .................... .................... ....................
F 7130 hCoV-19/South Korea/KCDC2458/2020|EPI_ISL_514861|2 .................... .................... .................... .................... .................... ....................
F 7131 hCoV-19/South Korea/KCDC2459/2020|EPI_ISL_514862|2 .................... .................... .................... .................... .................... ....................
F 7132 hCoV-19/South Korea/KCDC2460/2020|EPI_ISL_514863|2 .................... .................... .................... .................... .................... ....................
F 7133 hCoV-19/South Korea/KCDC2461/2020|EPI_ISL_514864|2 .................... .................... .................... .................... .................... ....................
F 7134 hCoV-19/South Korea/KCDC2462/2020|EPI_ISL_514865|2 .................... .................... .................... .................... .................... ....................
F 7135 hCoV-19/South Korea/KCDC2463/2020|EPI_ISL_514866|2 .................... .................... .................... .................... .................... ....................
F 7136 hCoV-19/South Korea/KCDC2464/2020|EPI_ISL_514867|2 .................... .................... .................... .................... .................... ....................
F 7137 hCoV-19/South Korea/KCDC2465/2020|EPI_ISL_514868|2 .................... .................... .................... .................... .................... ....................
F 7138 hCoV-19/South Korea/KCDC2466/2020|EPI_ISL_514869|2 .................... .................... .................... .................... .................... ....................
F 7139 hCoV-19/South Korea/KCDC2467/2020|EPI_ISL_514870|2 .................... .................... .................... .................... .................... ....................
F 7140 hCoV-19/South Korea/KCDC2468/2020|EPI_ISL_514871|2 .................... .................... .................... .................... .................... ....................
F 7141 hCoV-19/South Korea/KCDC2469/2020|EPI_ISL_514872|2 .................... .................... .................... .................... .................... ....................
F 7142 hCoV-19/South Korea/KCDC2470/2020|EPI_ISL_514873|2 .................... .................... .................... .................... .................... ....................
F 7143 hCoV-19/South Korea/KCDC2471/2020|EPI_ISL_514874|2 .................... .................... .................... .................... .................... ....................
F 7144 hCoV-19/South Korea/KCDC2472/2020|EPI_ISL_514875|2 .................... .................... .................... .................... .................... ....................
F 7145 hCoV-19/South Korea/KCDC2473/2020|EPI_ISL_514876|2 .................... .................... .................... .................... .................... ....................
F 7146 hCoV-19/South Korea/KCDC2474/2020|EPI_ISL_514877|2 .................... .................... .................... .................... .................... ....................
F 7147 hCoV-19/South Korea/KCDC2475/2020|EPI_ISL_514878|2 .................... .................... .................... .................... .................... ....................
F 7148 hCoV-19/South Korea/KCDC2476/2020|EPI_ISL_514879|2 .................... .................... .................... .................... .................... ....................
F 7149 hCoV-19/South Korea/KCDC2477/2020|EPI_ISL_514880|2 .................... .................... .................... .................... .................... ....................
F 7150 hCoV-19/South Korea/KCDC2478/2020|EPI_ISL_514881|2 .................... .................... .................... .................... .................... ....................
F 7151 hCoV-19/South Korea/KCDC2479/2020|EPI_ISL_514882|2 .................... .................... .................... .................... .................... ....................
F 7152 hCoV-19/South Korea/KCDC2480/2020|EPI_ISL_514883|2 .................... .................... .................... .................... .................... ....................
F 7153 hCoV-19/South Korea/KCDC2481/2020|EPI_ISL_514884|2 .................... .................... .................... .................... .................... ....................
F 7154 hCoV-19/South Korea/KCDC2482/2020|EPI_ISL_514885|2 .................... .................... .................... .................... .................... ....................
F 7155 hCoV-19/South Korea/KCDC2483/2020|EPI_ISL_514886|2 .................... .................... .................... .................... .................... ....................
F 7156 hCoV-19/South Korea/KCDC2484/2020|EPI_ISL_514887|2 .................... .................... .................... .................... .................... ....................
F 7157 hCoV-19/South Korea/KCDC2485/2020|EPI_ISL_514888|2 .................... .................... .................... .................... .................... ....................
F 7158 hCoV-19/South Korea/KCDC2486/2020|EPI_ISL_514889|2 .................... .................... .................... .................... .................... ....................
F 7159 hCoV-19/South Korea/KCDC2487/2020|EPI_ISL_514890|2 .................... .................... .................... .................... .................... ....................
F 7160 hCoV-19/South Korea/KCDC2488/2020|EPI_ISL_514891|2 .................... .................... .................... .................... .................... ....................
F 7161 hCoV-19/South Korea/KCDC2489/2020|EPI_ISL_514892|2 .................... .................... .................... .................... .................... ....................
F 7162 hCoV-19/South Korea/KCDC2490/2020|EPI_ISL_514893|2 .................... .................... .................... .................... .................... ....................
F 7163 hCoV-19/South Korea/KCDC2491/2020|EPI_ISL_514894|2 .................... .................... .................... .................... .................... ....................
F 7164 hCoV-19/South Korea/KCDC2492/2020|EPI_ISL_514895|2 .................... .................... .................... .................... .................... ....................
F 7165 hCoV-19/South Korea/KCDC2493/2020|EPI_ISL_514896|2 .................... .................... .................... .................... .................... ....................
F 7166 hCoV-19/South Korea/KCDC2494/2020|EPI_ISL_514897|2 .................... .................... .................... .................... .................... ....................
F 7167 hCoV-19/South Korea/KCDC2495/2020|EPI_ISL_514898|2 .................... .................... .................... .................... .................... ....................
F 7168 hCoV-19/South Korea/KCDC2496/2020|EPI_ISL_514899|2 .................... .................... .................... .................... .................... ....................
F 7169 hCoV-19/India/MH-AFMC-5306/2020|EPI_ISL_496602|202 .................... .................... .................... .................... .................... ....................
F 7170 hCoV-19/Zhejiang/SX0715/2020|EPI_ISL_497950|2020-0 .................... .................... .................... .................... .................... ....................
F 7171 hCoV-19/South Korea/KCDC2030/2020|EPI_ISL_497951|2 .................... .................... .................... .................... .................... ....................
F 7172 hCoV-19/South Korea/KCDC2032/2020|EPI_ISL_497953|2 .................... .................... .................... .................... .................... ....................
F 7173 hCoV-19/South Korea/KCDC2033/2020|EPI_ISL_497954|2 .................... .................... .................... .................... .................... ....................
F 7174 hCoV-19/South Korea/KCDC2034/2020|EPI_ISL_497955|2 .................... .................... .................... .................... .................... ....................
F 7175 hCoV-19/South Korea/KCDC2035/2020|EPI_ISL_497956|2 .................... .................... .................... .................... .................... ....................
F 7176 hCoV-19/South Korea/KCDC2036/2020|EPI_ISL_497957|2 .................... .................... .................... .................... .................... ....................
F 7177 hCoV-19/South Korea/KCDC2037/2020|EPI_ISL_497958|2 .................... .................... .................... .................... .................... ....................
F 7178 hCoV-19/South Korea/KCDC2038/2020|EPI_ISL_497959|2 .................... .................... .................... .................... .................... ....................
F 7179 hCoV-19/South Korea/KCDC2039/2020|EPI_ISL_497960|2 .................... .................... .................... .................... .................... ....................
F 7180 hCoV-19/South Korea/KCDC2060/2020|EPI_ISL_497961|2 .................... .................... .................... .................... .................... ....................
F 7181 hCoV-19/South Korea/KCDC2061/2020|EPI_ISL_497962|2 .................... .................... .................... .................... .................... ....................
F 7182 hCoV-19/South Korea/KCDC2062/2020|EPI_ISL_497963|2 .................... .................... .................... .................... .................... ....................
F 7183 hCoV-19/South Korea/KCDC2063/2020|EPI_ISL_497964|2 .................... .................... .................... .................... .................... ....................
F 7184 hCoV-19/South Korea/KCDC2064/2020|EPI_ISL_497965|2 .................... .................... .................... .................... .................... ....................
F 7185 hCoV-19/South Korea/KCDC2065/2020|EPI_ISL_497966|2 .................... .................... .................... .................... .................... ....................
F 7186 hCoV-19/South Korea/KCDC2066/2020|EPI_ISL_497967|2 .................... .................... .................... .................... .................... ....................
F 7187 hCoV-19/South Korea/KCDC2067/2020|EPI_ISL_497968|2 .................... .................... .................... .................... .................... ....................
F 7188 hCoV-19/South Korea/KCDC2068/2020|EPI_ISL_497969|2 .................... .................... .................... .................... .................... ....................
F 7189 hCoV-19/South Korea/KCDC2070/2020|EPI_ISL_497971|2 .................... .................... .................... .................... .................... ....................
F 7190 hCoV-19/South Korea/KCDC2075/2020|EPI_ISL_497976|2 .................... .................... .................... .................... .................... ....................
F 7191 hCoV-19/South Korea/KCDC2076/2020|EPI_ISL_497977|2 .................... .................... .................... .................... .................... ....................
F 7192 hCoV-19/South Korea/KCDC2077/2020|EPI_ISL_497978|2 .................... .................... .................... .................... .................... ....................
F 7193 hCoV-19/South Korea/KCDC2079/2020|EPI_ISL_497980|2 .................... .................... .................... .................... .................... ....................
F 7194 hCoV-19/South Korea/KCDC2080/2020|EPI_ISL_497981|2 .................... .................... .................... .................... .................... ....................
F 7195 hCoV-19/South Korea/KCDC2081/2020|EPI_ISL_497982|2 .................... .................... .................... .................... .................... ....................
F 7196 hCoV-19/South Korea/KCDC2082/2020|EPI_ISL_497983|2 .................... .................... .................... .................... .................... ....................
F 7197 hCoV-19/South Korea/KCDC2083/2020|EPI_ISL_497984|2 .................... .................... .................... .................... .................... ....................
F 7198 hCoV-19/South Korea/KCDC2084/2020|EPI_ISL_497985|2 .................... .................... .................... .................... .................... ....................
F 7199 hCoV-19/South Korea/KCDC2085/2020|EPI_ISL_497986|2 .................... .................... .................... .................... .....T.............. ....................
F 7200 hCoV-19/South Korea/KCDC2086/2020|EPI_ISL_497987|2 .................... .................... .................... .................... .................... ....................
F 7201 hCoV-19/South Korea/KCDC2087/2020|EPI_ISL_497988|2 .................... .................... .................... .................... .................... ....................
F 7202 hCoV-19/South Korea/KCDC2088/2020|EPI_ISL_497989|2 .................... .................... .................... .................... .................... ....................
F 7203 hCoV-19/South Korea/KCDC2089/2020|EPI_ISL_497990|2 .................... .................... .................... .................... .................... ....................
F 7204 hCoV-19/South Korea/KCDC2090/2020|EPI_ISL_497991|2 .................... .................... .................... .................... .................... ....................
F 7205 hCoV-19/South Korea/KCDC2091/2020|EPI_ISL_497992|2 .................... .................... .................... .................... .................... ....................
F 7206 hCoV-19/South Korea/KCDC2092/2020|EPI_ISL_497993|2 .................... .................... .................... .................... .................... ....................
F 7207 hCoV-19/South Korea/KCDC2093/2020|EPI_ISL_497994|2 .................... .................... .................... .................... .................... ....................
F 7208 hCoV-19/South Korea/KCDC2094/2020|EPI_ISL_497995|2 .................... .................... .................... .................... .................... ....................

| | | | | | | | | |
|---|---|---|---|---|---|---|---|---|
| F | 7209 | hCoV-19/South Korea/KCDC2095/2020\|EPI_ISL_497996\|2 | .................... | .................... | .................... | .................... | .................... | .................... |
| F | 7210 | hCoV-19/South Korea/KCDC2096/2020\|EPI_ISL_497997\|2 | .................... | .................... | .................... | .................... | .................... | .................... |
| F | 7211 | hCoV-19/South Korea/KCDC2097/2020\|EPI_ISL_497998\|2 | .................... | .................... | .................... | .................... | .................... | .................... |
| F | 7212 | hCoV-19/South Korea/KCDC2098/2020\|EPI_ISL_497999\|2 | .................... | .................... | .................... | .................... | .................... | .................... |
| F | 7213 | hCoV-19/South Africa/KRISP-0568/2020\|EPI_ISL_49810 | .................... | .................... | .................... | .................... | .................... | .................... |
| F | 7215 | hCoV-19/Belgium/ULG-10231/2020\|EPI_ISL_498137\|2020 | .................... | .................... | .................... | .................... | .................... | .................... |
| F | 7216 | hCoV-19/Belgium/ULG-10232/2020\|EPI_ISL_498138\|2020 | .................... | .................... | .................... | .................... | .................... | .................... |
| F | 7217 | hCoV-19/Belgium/ULG-10233/2020\|EPI_ISL_498139\|2020 | .................... | .................... | .................... | .................... | .................... | .................... |
| F | 7218 | hCoV-19/Belgium/ULG-10235/2020\|EPI_ISL_498141\|2020 | .................... | .................... | .................... | .................... | .................... | .................... |
| F | 7219 | hCoV-19/Belgium/ULG-10237/2020\|EPI_ISL_498143\|2020 | .................... | .................... | .................... | .................... | .................... | .................... |
| F | 7220 | hCoV-19/Belgium/ULG-10238/2020\|EPI_ISL_498144\|2020 | .................... | .................... | .................... | .................... | .................... | .................... |
| F | 7221 | hCoV-19/Belgium/ULG-10240/2020\|EPI_ISL_498146\|2020 | .................... | .................... | .................... | .................... | .................... | .................... |
| F | 7222 | hCoV-19/Belgium/ULG-10243/2020\|EPI_ISL_498149\|2020 | .................... | .................... | .................... | .................... | .................... | .................... |
| F | 7223 | hCoV-19/Belgium/ULG-10244/2020\|EPI_ISL_498150\|2020 | .................... | .................... | .................... | .................... | .................... | .................... |
| F | 7224 | hCoV-19/Belgium/ULG-10245/2020\|EPI_ISL_498151\|2020 | .................... | .................... | .................... | .................... | .................... | .................... |
| F | 7225 | hCoV-19/Colombia/MAG-INS-87968/2020\|EPI_ISL_498152 | .................... | .................... | .................... | .................... | .................... | .................... |
| F | 7226 | hCoV-19/Colombia/CES-INS-87973/2020\|EPI_ISL_498153 | .................... | .................... | .................... | .................... | .................... | .................... |
| F | 7227 | hCoV-19/Colombia/CUN-INS-88496/2020\|EPI_ISL_498154 | .................... | .................... | .................... | .................... | .................... | .................... |
| F | 7228 | hCoV-19/Colombia/ATL-INS-88612/2020\|EPI_ISL_498155 | .................... | .................... | .................... | .................... | .................... | .................... |
| F | 7229 | hCoV-19/Colombia/ATL-INS-88712/2020\|EPI_ISL_498156 | .................... | .................... | .................... | .................... | .................... | .................... |
| F | 7230 | hCoV-19/Colombia/BOY-INS-88859/2020\|EPI_ISL_498157 | .................... | .................... | .................... | .................... | .................... | .................... |
| F | 7231 | hCoV-19/Colombia/BOY-INS-88871/2020\|EPI_ISL_498158 | .................... | .................... | .................... | .................... | .................... | .................... |
| F | 7232 | hCoV-19/Colombia/CAU-INS-89002/2020\|EPI_ISL_498159 | .................... | .................... | .................... | .................... | .................... | .................... |
| F | 7233 | hCoV-19/Colombia/CES-INS-89781/2020\|EPI_ISL_498160 | .................... | .................... | .................... | .................... | .................... | .................... |
| F | 7234 | hCoV-19/Colombia/BOL-INS-94411/2020\|EPI_ISL_498161 | .................... | .................... | .................... | .................... | .................... | .................... |
| F | 7235 | hCoV-19/Colombia/NAR-INS-94679/2020\|EPI_ISL_498162 | .................... | .................... | .................... | .................... | .................... | .................... |
| F | 7236 | hCoV-19/Colombia/COR-INS-95319/2020\|EPI_ISL_498163 | .................... | .................... | .................... | .................... | .................... | .................... |
| F | 7237 | hCoV-19/Colombia/MET-INS-95637/2020\|EPI_ISL_498164 | .................... | .................... | .................... | .................... | .................... | .................... |
| F | 7238 | hCoV-19/Colombia/RIS-INS-95663/2020\|EPI_ISL_498165 | .................... | .................... | .................... | .................... | .................... | .................... |
| F | 7239 | hCoV-19/Colombia/MAG-INS-95839/2020\|EPI_ISL_498166 | .................... | .................... | .................... | .................... | .................... | .................... |
| F | 7240 | hCoV-19/Colombia/CUN-INS-96791/2020\|EPI_ISL_498167 | .................... | .................... | .................... | .................... | .................... | .................... |
| F | 7241 | hCoV-19/Colombia/CUN-INS-T1/2020\|EPI_ISL_498168\|20 | .................... | .................... | .................... | .................... | .................... | .................... |
| F | 7242 | hCoV-19/Colombia/CUN-INS-T8/2020\|EPI_ISL_498169\|20 | .................... | .................... | .................... | .................... | .................... | .................... |
| F | 7243 | hCoV-19/Colombia/CUN-INS-T12/2020\|EPI_ISL_498170\|2 | .................... | .................... | .................... | .................... | .................... | .................... |
| F | 7244 | hCoV-19/Vietnam/HCMC-35022/2020\|EPI_ISL_498171\|202 | .................... | .................... | .................... | .................... | .................... | .................... |
| F | 7245 | hCoV-19/Vietnam/HCMC-2712/2020\|EPI_ISL_498172\|2020 | .................... | .................... | .................... | .................... | .................... | .................... |
| F | 7246 | hCoV-19/Vietnam/HCMC-2751/2020\|EPI_ISL_498173\|2020 | .................... | .................... | .................... | .................... | .................... | .................... |
| F | 7247 | hCoV-19/Vietnam/HCMC-3013/2020\|EPI_ISL_498174\|2020 | .................... | .................... | .................... | .................... | .................... | .................... |
| F | 7248 | hCoV-19/Vietnam/HCMC-3349/2020\|EPI_ISL_498175\|2020 | .................... | .................... | .................... | .................... | .................... | .................... |
| F | 7249 | hCoV-19/Vietnam/HCMC-05003/2020\|EPI_ISL_498176\|202 | .................... | .................... | .................... | .................... | .................... | .................... |
| F | 7250 | hCoV-19/Vietnam/HCMC-05006/2020\|EPI_ISL_498177\|202 | .................... | .................... | .................... | .................... | .................... | .................... |
| F | 7251 | hCoV-19/Vietnam/HCMC-05007/2020\|EPI_ISL_498178\|202 | .................... | .................... | .................... | .................... | .................... | .................... |
| F | 7252 | hCoV-19/Vietnam/HCMC-05008/2020\|EPI_ISL_498179\|202 | .................... | .................... | .................... | .................... | .................... | .................... |
| F | 7253 | hCoV-19/Wales/PHWC-1693ED/2020\|EPI_ISL_499439\|2020 | .................... | .................... | .................... | .................... | .................... | .................... |
| F | 7254 | hCoV-19/England/LIVE-A692F/2020\|EPI_ISL_499468\|202 | .................... | .................... | .................... | .................... | .................... | .................... |
| F | 7255 | hCoV-19/England/LIVE-A6725/2020\|EPI_ISL_499469\|202 | .................... | .................... | .................... | .................... | .................... | .................... |
| F | 7257 | hCoV-19/Spain/PV-IBV-004230/2020\|EPI_ISL_500294\|20 | .................... | .................... | .................... | .................... | .................... | .................... |
| F | 7261 | hCoV-19/USA/MI-MDHHS-SC21446/2020\|EPI_ISL_507700\|2 | .................... | .................... | .................... | .................... | .................... | .................... |
| F | 7262 | hCoV-19/USA/MI-MDHHS-SC21447/2020\|EPI_ISL_507701\|2 | .................... | .................... | .................... | .................... | .................... | .................... |
| F | 7263 | hCoV-19/USA/MI-MDHHS-SC21448/2020\|EPI_ISL_507702\|2 | .................... | .................... | .................... | .................... | .................... | .................... |
| F | 7264 | hCoV-19/USA/MI-MDHHS-SC21449/2020\|EPI_ISL_507703\|2 | .................... | .................... | .................... | .................... | .................... | .................... |
| F | 7265 | hCoV-19/USA/MI-MDHHS-SC21450/2020\|EPI_ISL_507704\|2 | .................... | .................... | .................... | .................... | .................... | .................... |
| F | 7266 | hCoV-19/USA/MI-MDHHS-SC21451/2020\|EPI_ISL_507705\|2 | .................... | .................... | .................... | .................... | .................... | .................... |
| F | 7267 | hCoV-19/USA/MI-MDHHS-SC21452/2020\|EPI_ISL_507706\|2 | .................... | .................... | .................... | .................... | .................... | .................... |
| F | 7268 | hCoV-19/USA/MI-MDHHS-SC21453/2020\|EPI_ISL_507707\|2 | .................... | .................... | .................... | .................... | .................... | .................... |
| F | 7269 | hCoV-19/USA/MI-MDHHS-SC21454/2020\|EPI_ISL_507708\|2 | .................... | .................... | .................... | .................... | .................... | .................... |
| F | 7270 | hCoV-19/USA/MI-MDHHS-SC21455/2020\|EPI_ISL_507709\|2 | .................... | .................... | .................... | .................... | .................... | .................... |
| F | 7271 | hCoV-19/USA/MI-MDHHS-SC21456/2020\|EPI_ISL_507710\|2 | .................... | .................... | .................... | .................... | .................... | .................... |
| F | 7272 | hCoV-19/USA/MI-MDHHS-SC21457/2020\|EPI_ISL_507711\|2 | .................... | .................... | .................... | .................... | .................... | .................... |
| F | 7273 | hCoV-19/USA/MI-MDHHS-SC21458/2020\|EPI_ISL_507712\|2 | .................... | .................... | .................... | .................... | .................... | .................... |
| F | 7274 | hCoV-19/USA/MI-MDHHS-SC21459/2020\|EPI_ISL_507713\|2 | .................... | .................... | .................... | .................... | .................... | .................... |
| F | 7275 | hCoV-19/USA/MI-MDHHS-SC21460/2020\|EPI_ISL_507714\|2 | .................... | .................... | .................... | .................... | .................... | .................... |
| F | 7276 | hCoV-19/USA/MI-MDHHS-SC21461/2020\|EPI_ISL_507715\|2 | .................... | .................... | .................... | .................... | .................... | .................... |
| F | 7277 | hCoV-19/USA/MI-MDHHS-SC21462/2020\|EPI_ISL_507716\|2 | .................... | .................... | .................... | .................... | .................... | .................... |
| F | 7278 | hCoV-19/USA/MI-MDHHS-SC21463/2020\|EPI_ISL_507717\|2 | .................... | .................... | .................... | .................... | .................... | .................... |
| F | 7279 | hCoV-19/USA/MI-MDHHS-SC21464/2020\|EPI_ISL_507718\|2 | .................... | .................... | .................... | .................... | .................... | .................... |
| F | 7280 | hCoV-19/USA/MI-MDHHS-SC21465/2020\|EPI_ISL_507719\|2 | .................... | .................... | .................... | .................... | .................... | .................... |
| F | 7281 | hCoV-19/USA/MI-MDHHS-SC21466/2020\|EPI_ISL_507720\|2 | .................... | .................... | .................... | .................... | .................... | .................... |
| F | 7290 | hCoV-19/USA/MI-MDHHS-SC20991/2020\|EPI_ISL_507777\|2 | .................... | .................... | .................... | .................... | .................... | .................... |
| F | 7291 | hCoV-19/USA/MI-MDHHS-SC20992/2020\|EPI_ISL_507778\|2 | .................... | .................... | .................... | .................... | .................... | .................... |
| F | 7292 | hCoV-19/USA/MI-MDHHS-SC20993/2020\|EPI_ISL_507779\|2 | .................... | .................... | .................... | .................... | .................... | .................... |
| F | 7293 | hCoV-19/USA/MI-MDHHS-SC20994/2020\|EPI_ISL_507780\|2 | .................... | .................... | .................... | .................... | .................... | .................... |
| F | 7294 | hCoV-19/USA/MI-MDHHS-SC20995/2020\|EPI_ISL_507781\|2 | .................... | .................... | .................... | .................... | .................... | .................... |
| F | 7295 | hCoV-19/USA/MI-MDHHS-SC20996/2020\|EPI_ISL_507782\|2 | .................... | .................... | .................... | .................... | .................... | .................... |
| F | 7296 | hCoV-19/USA/MI-MDHHS-SC20997/2020\|EPI_ISL_507783\|2 | .................... | .................... | .................... | .................... | .................... | .................... |
| F | 7297 | hCoV-19/USA/MI-MDHHS-SC20998/2020\|EPI_ISL_507784\|2 | .................... | .................... | .................... | .................... | .................... | .................... |
| F | 7298 | hCoV-19/USA/MI-MDHHS-SC20999/2020\|EPI_ISL_507785\|2 | .................... | .................... | .................... | .................... | .................... | .................... |
| F | 7299 | hCoV-19/USA/MI-MDHHS-SC21000/2020\|EPI_ISL_507786\|2 | .................... | .................... | .................... | .................... | .................... | .................... |
| F | 7300 | hCoV-19/USA/MI-MDHHS-SC21001/2020\|EPI_ISL_507787\|2 | .................... | .................... | .................... | .................... | .................... | .................... |
| F | 7301 | hCoV-19/USA/MI-MDHHS-SC21002/2020\|EPI_ISL_507788\|2 | .................... | .................... | .................... | .................... | .................... | .................... |

| | | | | | | | |
|---|---|---|---|---|---|---|---|
| F | 7302 hCoV-19/USA/MI-MDHHS-SC21003/2020\|EPI_ISL_507789\|2 | ···················· | ···················· | ···················· | ···················· | ···················· | ···················· |
| F | 7303 hCoV-19/USA/NM-UNM-00374/2020\|EPI_ISL_508013\|2020- | ···················· | ···················· | ···················· | ···················· | ···················· | ···················· |
| F | 7304 hCoV-19/USA/NM-UNM-00281/2020\|EPI_ISL_508014\|2020- | ···················· | ···················· | ···················· | ···················· | ···················· | ···················· |
| F | 7305 hCoV-19/USA/NM-UNM-00334/2020\|EPI_ISL_508018\|2020- | ···················· | ···················· | ···················· | ···················· | ···················· | ···················· |
| F | 7306 hCoV-19/USA/NM-UNM-00335/2020\|EPI_ISL_508026\|2020- | ···················· | ···················· | ···················· | ···················· | ···················· | ···················· |
| F | 7307 hCoV-19/USA/NM-UNM-00278/2020\|EPI_ISL_508029\|2020- | ···················· | ···················· | ···················· | ···················· | ···················· | ···················· |
| F | 7308 hCoV-19/South Africa/KRISP-K001075/2020\|EPI_ISL_50 | ···················· | ···················· | ···················· | ···················· | ···················· | ···················· |
| F | 7309 hCoV-19/South Africa/KRISP-K001088/2020\|EPI_ISL_50 | ···················· | ···················· | ···················· | ···················· | ···················· | ···················· |
| F | 7310 hCoV-19/South Africa/KRISP-K001089/2020\|EPI_ISL_50 | ···················· | ···················· | ···················· | ···················· | ···················· | ···················· |
| F | 7311 hCoV-19/South Africa/KRISP-K001091/2020\|EPI_ISL_50 | ···················· | ···················· | ···················· | ···················· | ···················· | ···················· |
| F | 7312 hCoV-19/South Africa/KRISP-K001092/2020\|EPI_ISL_50 | ···················· | ···················· | ···················· | ···················· | ···················· | ···················· |
| F | 7313 hCoV-19/South Africa/KRISP-K001093/2020\|EPI_ISL_50 | ···················· | ···················· | ···················· | ···················· | ···················· | ···················· |
| F | 7314 hCoV-19/South Africa/KRISP-K001096/2020\|EPI_ISL_50 | ···················· | ···················· | ···················· | ···················· | ···················· | ···················· |
| F | 7315 hCoV-19/South Africa/KRISP-K001097/2020\|EPI_ISL_50 | ···················· | ···················· | ···················· | ···················· | ···················· | ···················· |
| F | 7316 hCoV-19/South Africa/KRISP-K001098/2020\|EPI_ISL_50 | ···················· | ···················· | ···················· | ···················· | ···················· | ···················· |
| F | 7317 hCoV-19/South Africa/KRISP-K001099/2020\|EPI_ISL_50 | ···················· | ···················· | ···················· | ···················· | ···················· | ···················· |
| F | 7318 hCoV-19/South Africa/KRISP-K001100/2020\|EPI_ISL_50 | ···················· | ···················· | ···················· | ···················· | ···················· | ···················· |
| F | 7319 hCoV-19/South Africa/KRISP-K001101/2020\|EPI_ISL_50 | ···················· | ···················· | ···················· | ···················· | ···················· | ···················· |
| F | 7320 hCoV-19/South Africa/KRISP-K001102/2020\|EPI_ISL_50 | ···················· | ···················· | ···················· | ···················· | ···················· | ···················· |
| F | 7321 hCoV-19/South Africa/KRISP-K001103/2020\|EPI_ISL_50 | ···················· | ···················· | ···················· | ···················· | ···················· | ···················· |
| F | 7322 hCoV-19/South Africa/KRISP-K001104/2020\|EPI_ISL_50 | ···················· | ···················· | ···················· | ···················· | ···················· | ···················· |
| F | 7323 hCoV-19/South Africa/KRISP-K001105/2020\|EPI_ISL_50 | ···················· | ···················· | ···················· | ···················· | ···················· | ···················· |
| F | 7324 hCoV-19/South Africa/KRISP-K001106/2020\|EPI_ISL_50 | ···················· | ···················· | ···················· | ···················· | ···················· | ···················· |
| F | 7325 hCoV-19/South Africa/KRISP-K001109/2020\|EPI_ISL_50 | ···················· | ···················· | ···················· | ···················· | ···················· | ···················· |
| F | 7326 hCoV-19/South Africa/KRISP-K001110/2020\|EPI_ISL_50 | ···················· | ···················· | ···················· | ···················· | ···················· | ···················· |
| F | 7327 hCoV-19/South Africa/KRISP-K001112/2020\|EPI_ISL_50 | ···················· | ···················· | ···················· | ···················· | ···················· | ···················· |
| F | 7328 hCoV-19/South Africa/KRISP-K001114/2020\|EPI_ISL_50 | ···················· | ···················· | ···················· | ···················· | ···················· | ···················· |
| F | 7329 hCoV-19/South Africa/KRISP-K001115/2020\|EPI_ISL_50 | ···················· | ···················· | ···················· | ···················· | ···················· | ···················· |
| F | 7330 hCoV-19/South Africa/KRISP-K001116/2020\|EPI_ISL_50 | ···················· | ···················· | ···················· | ···················· | ···················· | ···················· |
| F | 7331 hCoV-19/South Africa/KRISP-K001117/2020\|EPI_ISL_50 | ···················· | ···················· | ···················· | ···················· | ···················· | ···················· |
| F | 7332 hCoV-19/South Africa/KRISP-K001118/2020\|EPI_ISL_50 | ···················· | ···················· | ···················· | ···················· | ···················· | ···················· |
| F | 7333 hCoV-19/South Africa/KRISP-K001121/2020\|EPI_ISL_50 | ···················· | ···················· | ···················· | ···················· | ···················· | ···················· |
| F | 7334 hCoV-19/South Africa/KRISP-K001123/2020\|EPI_ISL_50 | ···················· | ···················· | ···················· | ···················· | ···················· | ···················· |
| F | 7335 hCoV-19/South Africa/KRISP-K001124/2020\|EPI_ISL_50 | ···················· | ···················· | ···················· | ···················· | ···················· | ···················· |
| F | 7336 hCoV-19/South Africa/KRISP-K001125/2020\|EPI_ISL_50 | ···················· | ···················· | ···················· | ···················· | ···················· | ···················· |
| F | 7337 hCoV-19/South Africa/KRISP-K001126/2020\|EPI_ISL_50 | ···················· | ···················· | ···················· | ···················· | ···················· | ···················· |
| F | 7338 hCoV-19/Singapore/763/2020\|EPI_ISL_509373\|2020-06- | ···················· | ···················· | ···················· | ···················· | ···················· | ···················· |
| F | 7339 hCoV-19/Singapore/764/2020\|EPI_ISL_509374\|2020-06- | ···················· | ···················· | ···················· | ···················· | ···················· | ···················· |
| F | 7340 hCoV-19/Singapore/765/2020\|EPI_ISL_509375\|2020-06- | ···················· | ···················· | ···················· | ···················· | ···················· | ···················· |
| F | 7341 hCoV-19/Singapore/767/2020\|EPI_ISL_509377\|2020-06- | ···················· | ···················· | ···················· | ···················· | ···················· | ···················· |
| F | 7342 hCoV-19/Singapore/768/2020\|EPI_ISL_509378\|2020-06- | ···················· | ···················· | ···················· | ···················· | ···················· | ···················· |
| F | 7343 hCoV-19/Singapore/769/2020\|EPI_ISL_509379\|2020-07- | ···················· | ···················· | ···················· | ···················· | ···················· | ···················· |
| F | 7349 hCoV-19/Spain/EX-IBV-005019/2020\|EPI_ISL_510328\|20 | ···················· | ···················· | ···················· | ···················· | ···················· | ···················· |
| F | 7352 hCoV-19/Spain/EX-IBV-005022/2020\|EPI_ISL_510331\|20 | ···················· | ···················· | ···················· | ···················· | ···················· | ···················· |
| F | 7364 hCoV-19/Spain/AR-IBV-005067/2020\|EPI_ISL_510357\|20 | ···················· | ···················· | ···················· | ···················· | ···················· | ···················· |
| F | 7368 hCoV-19/Spain/AR-IBV-005075/2020\|EPI_ISL_510361\|20 | ···················· | ···················· | ···················· | ···················· | ··N················· | ···················· |
| F | 7372 hCoV-19/Portugal/PT1102/2020\|EPI_ISL_511649\|2020-0 | ···················· | ···················· | ···················· | ···················· | ···················· | ···················· |
| F | 7373 hCoV-19/Portugal/PT1109/2020\|EPI_ISL_511656\|2020-0 | ···················· | ···················· | ···················· | ···················· | ···················· | ···················· |
| F | 7374 hCoV-19/Portugal/PT1110/2020\|EPI_ISL_511657\|2020-0 | ···················· | ···················· | ···················· | ···················· | ···················· | ···················· |
| F | 7375 hCoV-19/Portugal/PT1111/2020\|EPI_ISL_511658\|2020-0 | ···················· | ···················· | ···················· | ···················· | ···················· | ···················· |
| F | 7376 hCoV-19/Portugal/PT1112/2020\|EPI_ISL_511659\|2020-0 | ···················· | ···················· | ···················· | ···················· | ···················· | ···················· |
| F | 7377 hCoV-19/Portugal/PT1113/2020\|EPI_ISL_511660\|2020-0 | ···················· | ···················· | ···················· | ···················· | ···················· | ···················· |
| F | 7378 hCoV-19/Portugal/PT1114/2020\|EPI_ISL_511661\|2020-0 | ···················· | ···················· | ···················· | ···················· | ···················· | ···················· |
| F | 7379 hCoV-19/Portugal/PT1116/2020\|EPI_ISL_511663\|2020-0 | ···················· | ···················· | ···················· | ···················· | ···················· | ···················· |
| F | 7380 hCoV-19/Portugal/PT1121/2020\|EPI_ISL_511668\|2020-0 | ···················· | ···················· | ···················· | ···················· | ···················· | ···················· |
| F | 7381 hCoV-19/Portugal/PT1122/2020\|EPI_ISL_511669\|2020-0 | ···················· | ···················· | ···················· | ···················· | ···················· | ···················· |
| F | 7382 hCoV-19/Portugal/PT1123/2020\|EPI_ISL_511670\|2020-0 | ···················· | ···················· | ···················· | ···················· | ···················· | ···················· |
| F | 7383 hCoV-19/Portugal/PT1124/2020\|EPI_ISL_511671\|2020-0 | ···················· | ···················· | ···················· | ···················· | ···················· | ···················· |
| F | 7384 hCoV-19/Portugal/PT1125/2020\|EPI_ISL_511672\|2020-0 | ···················· | ···················· | ···················· | ···················· | ···················· | ···················· |
| F | 7385 hCoV-19/Portugal/PT1126/2020\|EPI_ISL_511673\|2020-0 | ···················· | ···················· | ···················· | ···················· | ···················· | ···················· |
| F | 7386 hCoV-19/Portugal/PT1127/2020\|EPI_ISL_511674\|2020-0 | ···················· | ···················· | ···················· | ···················· | ···················· | ···················· |
| F | 7387 hCoV-19/Portugal/PT1128/2020\|EPI_ISL_511675\|2020-0 | ···················· | ···················· | ···················· | ···················· | ···················· | ···················· |
| F | 7388 hCoV-19/Portugal/PT1129/2020\|EPI_ISL_511676\|2020-0 | ···················· | ···················· | ···················· | ···················· | ···················· | ···················· |
| F | 7389 hCoV-19/Portugal/PT1130/2020\|EPI_ISL_511677\|2020-0 | ···················· | ···················· | ···················· | ···················· | ···················· | ···················· |
| F | 7390 hCoV-19/Portugal/PT1131/2020\|EPI_ISL_511678\|2020-0 | ···················· | ···················· | ···················· | ···················· | ···················· | ···················· |
| F | 7391 hCoV-19/Portugal/PT1132/2020\|EPI_ISL_511679\|2020-0 | ···················· | ···················· | ···················· | ···················· | ···················· | ···················· |
| F | 7392 hCoV-19/Portugal/PT1133/2020\|EPI_ISL_511680\|2020-0 | ···················· | ···················· | ···················· | ···················· | ···················· | ···················· |
| F | 7393 hCoV-19/Portugal/PT1136/2020\|EPI_ISL_511683\|2020-0 | ···················· | ···················· | ···················· | ···················· | ···················· | ···················· |
| F | 7394 hCoV-19/Portugal/PT1137/2020\|EPI_ISL_511684\|2020-0 | ···················· | ···················· | ···················· | ···················· | ···················· | ···················· |
| F | 7395 hCoV-19/Portugal/PT1138/2020\|EPI_ISL_511685\|2020-0 | ···················· | ···················· | ···················· | ···················· | ···················· | ···················· |
| F | 7396 hCoV-19/Portugal/PT1140/2020\|EPI_ISL_511687\|2020-0 | ···················· | ···················· | ···················· | ···················· | ···················· | ···················· |
| F | 7397 hCoV-19/Portugal/PT1141/2020\|EPI_ISL_511688\|2020-0 | ···················· | ···················· | ···················· | ···················· | ···················· | ···················· |
| F | 7398 hCoV-19/Portugal/PT1142/2020\|EPI_ISL_511689\|2020-0 | ···················· | ···················· | ···················· | ···················· | ···················· | ···················· |
| F | 7399 hCoV-19/Portugal/PT1144/2020\|EPI_ISL_511691\|2020-0 | ···················· | ···················· | ···················· | ···················· | ···················· | ···················· |
| F | 7400 hCoV-19/Portugal/PT1145/2020\|EPI_ISL_511692\|2020-0 | ···················· | ···················· | ···················· | ···················· | ···················· | ···················· |
| F | 7401 hCoV-19/Portugal/PT1146/2020\|EPI_ISL_511693\|2020-0 | ···················· | ···················· | ···················· | ···················· | ···················· | ···················· |
| F | 7402 hCoV-19/Portugal/PT1148/2020\|EPI_ISL_511695\|2020-0 | ···················· | ···················· | ···················· | ···················· | ···················· | ···················· |
| F | 7403 hCoV-19/Portugal/PT1149/2020\|EPI_ISL_511696\|2020-0 | ···················· | ···················· | ···················· | ···················· | ···················· | ···················· |
| F | 7404 hCoV-19/USA/CA-ALSR-1972/2020\|EPI_ISL_512270\|2020- | ···················· | ·A·················· | ···················· | ···················· | ···················· | ···················· |
| F | 7405 hCoV-19/USA/CA-ALSR-1973/2020\|EPI_ISL_512271\|2020- | ···················· | ·A·················· | ···················· | ···················· | ···················· | ···················· |

| | | | | | | | | |
|---|---|---|---|---|---|---|---|---|
| F | 7406 | hCoV-19/USA/CA-ALSR-1975/2020\|EPI_ISL_512273\|2020- | ···················· | ·A·················· | ···················· | ···················· | ···················· | ···················· |
| F | 7407 | hCoV-19/Poland/1132916/2020\|EPI_ISL_512296\|2020-05 | ···················· | ···················· | ···················· | ···················· | ···················· | ···················· |
| F | 7408 | hCoV-19/Poland/1134056/2020\|EPI_ISL_512297\|2020-05 | ···················· | ···················· | ···················· | ···················· | ···················· | ···················· |
| F | 7409 | hCoV-19/Poland/1140683/2020\|EPI_ISL_512298\|2020-05 | ···················· | ···················· | ···················· | ···················· | ···················· | ···················· |
| F | 7410 | hCoV-19/Poland/1154942/2020\|EPI_ISL_512299\|2020-06 | ···················· | ···················· | ···················· | ···················· | ···················· | ···················· |
| F | 7411 | hCoV-19/SaudiArabia/KAUST-JEDDAH615/2020\|EPI_ISL_5 | ···················· | ···················· | ···················· | ···················· | ···················· | ···················· |
| F | 7412 | hCoV-19/Saudi Arabia/KAUST-JEDDAH632/2020\|EPI_ISL_ | ···················· | ···················· | ···················· | ···················· | ···················· | ···················· |
| F | 7413 | hCoV-19/SaudiArabia/KAUST-JEDDAH720/2020\|EPI_ISL_5 | ···················· | ···················· | ···················· | ···················· | ···················· | ···················· |
| F | 7414 | hCoV-19/SaudiArabia/KAUST-JEDDAH721/2020\|EPI_ISL_5 | ···················· | ···················· | ···················· | ···················· | ···················· | ···················· |
| F | 7415 | hCoV-19/Saudi Arabia/KAUST-JEDDAH722/2020\|EPI_ISL_ | ···················· | ···················· | ···················· | ···················· | ···················· | ···················· |
| F | 7416 | hCoV-19/Saudi Arabia/KAUST-JEDDAH723/2020\|EPI_ISL_ | ···················· | ···················· | ···················· | ···················· | ···················· | ···················· |
| F | 7417 | hCoV-19/SaudiArabia/KAUST-JEDDAH743/2020\|EPI_ISL_5 | ···················· | ···················· | ···················· | ···················· | ···················· | ···················· |
| F | 7418 | hCoV-19/SaudiArabia/KAUST-JEDDAH746/2020\|EPI_ISL_5 | ···················· | ···················· | ···················· | ···················· | ···················· | ···················· |
| F | 7419 | hCoV-19/SaudiArabia/KAUST-JEDDAH747/2020\|EPI_ISL_5 | ···················· | ···················· | ···················· | ···················· | ···················· | ···················· |
| F | 7420 | hCoV-19/SaudiArabia/KAUST-JEDDAH748/2020\|EPI_ISL_5 | ···················· | ···················· | ···················· | ···················· | ···················· | ···················· |
| F | 7421 | hCoV-19/SaudiArabia/KAUST-JEDDAH754/2020\|EPI_ISL_5 | ···················· | ···················· | ···················· | ···················· | ···················· | ···················· |
| F | 7422 | hCoV-19/SaudiArabia/KAUST-JEDDAH755/2020\|EPI_ISL_5 | ···················· | ·A·················· | ···················· | ···················· | ···················· | ···················· |
| F | 7423 | hCoV-19/SaudiArabia/KAUST-JEDDAH756/2020\|EPI_ISL_5 | ···················· | ···················· | ···················· | ···················· | ···················· | ···················· |
| F | 7424 | hCoV-19/Brazil/RJ-DCVN2/2020\|EPI_ISL_529139\|2020-0 | ···················· | ···················· | ···················· | ···················· | ···················· | ···················· |
| F | 7425 | hCoV-19/South Africa/KRISP-0561/2020\|EPI_ISL_49810 | ···················· | ···················· | ···················· | ···················· | ···················· | ···················· |
| F | 7426 | hCoV-19/South Africa/KRISP-0562/2020\|EPI_ISL_49810 | ···················· | ···················· | ···················· | ···················· | ···················· | ···················· |
| F | 7427 | hCoV-19/South Africa/KRISP-0563/2020\|EPI_ISL_49810 | ···················· | ···················· | ···················· | ···················· | ···················· | ···················· |
| F | 7428 | hCoV-19/South Africa/KRISP-0564/2020\|EPI_ISL_49810 | ···················· | ···················· | ···················· | ···················· | ···················· | ···················· |
| F | 7429 | hCoV-19/South Africa/KRISP-0565/2020\|EPI_ISL_49810 | ···················· | ···················· | ···················· | ···················· | ···················· | ···················· |
| F | 7431 | hCoV-19/South Africa/KRISP-0570/2020\|EPI_ISL_49810 | ···················· | ···················· | ···················· | ···················· | ···················· | ···················· |
| F | 7432 | hCoV-19/South Africa/KRISP-0571/2020\|EPI_ISL_49810 | ···················· | ···················· | ···················· | ···················· | ···················· | ···················· |
| F | 7433 | hCoV-19/South Africa/KRISP-0572/2020\|EPI_ISL_49811 | ···················· | ···················· | ···················· | ···················· | ···················· | ···················· |
| F | 7434 | hCoV-19/South Africa/KRISP-0573/2020\|EPI_ISL_49811 | ···················· | ···················· | ···················· | ···················· | ···················· | ···················· |
| F | 7435 | hCoV-19/South Africa/KRISP-0574/2020\|EPI_ISL_49811 | ···················· | ···················· | ···················· | ···················· | ···················· | ···················· |
| F | 7436 | hCoV-19/South Africa/KRISP-0575/2020\|EPI_ISL_49811 | ···················· | ···················· | ···················· | ···················· | ···················· | ···················· |
| F | 7437 | hCoV-19/South Africa/KRISP-0576/2020\|EPI_ISL_49811 | ···················· | ···················· | ···················· | ···················· | ···················· | ···················· |
| F | 7438 | hCoV-19/South Africa/KRISP-0577/2020\|EPI_ISL_49811 | ···················· | ···················· | ···················· | ···················· | ···················· | ···················· |
| F | 7439 | hCoV-19/South Africa/KRISP-0578/2020\|EPI_ISL_49811 | ···················· | ···················· | ···················· | ···················· | ···················· | ···················· |
| F | 7440 | hCoV-19/South Africa/KRISP-0579/2020\|EPI_ISL_49811 | ···················· | ···················· | ···················· | ···················· | ···················· | ···················· |
| F | 7441 | hCoV-19/South Africa/KRISP-0580/2020\|EPI_ISL_49811 | ···················· | ···················· | ···················· | ···················· | ···················· | ···················· |
| F | 7442 | hCoV-19/South Africa/KRISP-0581/2020\|EPI_ISL_49811 | ···················· | ···················· | ···················· | ···················· | ···················· | ···················· |
| F | 7443 | hCoV-19/South Africa/KRISP-0582/2020\|EPI_ISL_49812 | ···················· | ···················· | ···················· | ···················· | ···················· | ···················· |
| F | 7444 | hCoV-19/South Africa/KRISP-0583/2020\|EPI_ISL_49812 | ···················· | ···················· | ···················· | ···················· | ···················· | ···················· |
| F | 7445 | hCoV-19/South Africa/KRISP-0584/2020\|EPI_ISL_49812 | ···················· | ···················· | ···················· | ···················· | ···················· | ···················· |
| F | 7446 | hCoV-19/South Africa/KRISP-0585/2020\|EPI_ISL_49812 | ···················· | ···················· | ···················· | ···················· | ···················· | ···················· |
| F | 7447 | hCoV-19/South Africa/KRISP-0586/2020\|EPI_ISL_49812 | ···················· | ···················· | ···················· | ···················· | ···················· | ···················· |
| F | 7448 | hCoV-19/South Africa/KRISP-0587/2020\|EPI_ISL_49812 | ···················· | ···················· | ···················· | ···················· | ···················· | ···················· |
| F | 7449 | hCoV-19/South Africa/KRISP-0588/2020\|EPI_ISL_49812 | ···················· | ···················· | ···················· | ···················· | ···················· | ···················· |
| F | 7450 | hCoV-19/Belgium/ULG-10221/2020\|EPI_ISL_498127\|2020 | ···················· | ···················· | ···················· | ···················· | ···················· | ···················· |
| F | 7451 | hCoV-19/Belgium/ULG-10222/2020\|EPI_ISL_498128\|2020 | ···················· | ···················· | ···················· | ···················· | ···················· | ···················· |
| F | 7452 | hCoV-19/Belgium/ULG-10223/2020\|EPI_ISL_498129\|2020 | ···················· | ···················· | ···················· | ···················· | ···················· | ···················· |
| F | 7453 | hCoV-19/Belgium/ULG-10224/2020\|EPI_ISL_498130\|2020 | ···················· | ···················· | ···················· | ···················· | ···················· | ···················· |
| F | 7454 | hCoV-19/Belgium/ULG-10225/2020\|EPI_ISL_498131\|2020 | ···················· | ···················· | ···················· | ···················· | ···················· | ···················· |
| F | 7455 | hCoV-19/Belgium/ULG-10226/2020\|EPI_ISL_498132\|2020 | ···················· | ···················· | ···················· | ···················· | ···················· | ···················· |
| F | 7456 | hCoV-19/Belgium/ULG-10227/2020\|EPI_ISL_498133\|2020 | ···················· | ···················· | ···················· | ···················· | ···················· | ···················· |
| F | 7457 | hCoV-19/Belgium/ULG-10228/2020\|EPI_ISL_498134\|2020 | ···················· | ···················· | ···················· | ···················· | ···················· | ···················· |
| F | 7458 | hCoV-19/Belgium/ULG-10229/2020\|EPI_ISL_498135\|2020 | ···················· | ···················· | ···················· | ···················· | ·T·················· | ···················· |
| F | 7459 | hCoV-19/Belgium/ULG-10230/2020\|EPI_ISL_498136\|2020 | ···················· | ···················· | ···················· | ···················· | ···················· | ···················· |
| F | 7460 | hCoV-19/Wales/PHWC-169268/2020\|EPI_ISL_499400\|2020 | ···················· | ···················· | ···················· | ···················· | ···················· | ···················· |
| F | 7461 | hCoV-19/Wales/PHWC-1694DB/2020\|EPI_ISL_499401\|2020 | ···················· | ···················· | ···················· | ···················· | ···················· | ···················· |
| F | 7462 | hCoV-19/Wales/PHWC-1691C5/2020\|EPI_ISL_499417\|2020 | ···················· | ···················· | ···················· | ···················· | ···················· | ···················· |
| F | 7463 | hCoV-19/Wales/PHWC-169277/2020\|EPI_ISL_499424\|2020 | ···················· | ···················· | ···················· | ···················· | ···················· | ···················· |
| F | 7464 | hCoV-19/Wales/PHWC-169295/2020\|EPI_ISL_499425\|2020 | ···················· | ···················· | ···················· | ···················· | ···················· | ···················· |
| F | 7465 | hCoV-19/Wales/PHWC-1692A4/2020\|EPI_ISL_499426\|2020 | ···················· | ···················· | ···················· | ···················· | ···················· | ···················· |
| F | 7467 | hCoV-19/Wales/PHWC-1692FF/2020\|EPI_ISL_499428\|2020 | ···················· | ···················· | ···················· | ···················· | ···················· | ···················· |
| F | 7468 | hCoV-19/Wales/PHWC-16930B/2020\|EPI_ISL_499429\|2020 | ···················· | ···················· | ···················· | ···················· | ···················· | ···················· |
| F | 7469 | hCoV-19/Wales/PHWC-16931A/2020\|EPI_ISL_499430\|2020 | ···················· | ···················· | ···················· | ···················· | ···················· | ···················· |
| F | 7470 | hCoV-19/Wales/PHWC-169329/2020\|EPI_ISL_499431\|2020 | ···················· | ···················· | ···················· | ···················· | ···················· | ···················· |
| F | 7471 | hCoV-19/Wales/PHWC-169338/2020\|EPI_ISL_499432\|2020 | ···················· | ···················· | ···················· | ···················· | ···················· | ···················· |
| F | 7472 | hCoV-19/Wales/PHWC-169356/2020\|EPI_ISL_499433\|2020 | ···················· | ···················· | ···················· | ···················· | ···················· | ···················· |
| F | 7473 | hCoV-19/Wales/PHWC-169365/2020\|EPI_ISL_499434\|2020 | ···················· | ···················· | ···················· | ···················· | ···················· | ···················· |
| F | 7474 | hCoV-19/Wales/PHWC-169383/2020\|EPI_ISL_499435\|2020 | ···················· | ···················· | ···················· | ···················· | ···················· | ···················· |
| F | 7475 | hCoV-19/Wales/PHWC-169392/2020\|EPI_ISL_499436\|2020 | ···················· | ···················· | ···················· | ···················· | ···················· | ···················· |
| F | 7476 | hCoV-19/Wales/PHWC-1693B0/2020\|EPI_ISL_499437\|2020 | ···················· | ···················· | ···················· | ···················· | ···················· | ···················· |
| F | 7477 | hCoV-19/Wales/PHWC-1693DE/2020\|EPI_ISL_499438\|2020 | ···················· | ···················· | ···················· | ···················· | ···················· | ···················· |
| F | 7478 | hCoV-19/Wales/PHWC-1693FC/2020\|EPI_ISL_499440\|2020 | ···················· | ···················· | ···················· | ···················· | ···················· | ···················· |
| F | 7479 | hCoV-19/Wales/PHWC-169408/2020\|EPI_ISL_499441\|2020 | ···················· | ···················· | ···················· | ···················· | ···················· | ···················· |
| F | 7480 | hCoV-19/Wales/PHWC-169426/2020\|EPI_ISL_499442\|2020 | ···················· | ···················· | ···················· | ···················· | ···················· | ···················· |
| F | 7481 | hCoV-19/Wales/PHWC-169435/2020\|EPI_ISL_499443\|2020 | ···················· | ···················· | ···················· | ···················· | ···················· | ···················· |
| F | 7482 | hCoV-19/Wales/PHWC-169444/2020\|EPI_ISL_499444\|2020 | ···················· | ···················· | ···················· | ···················· | ···················· | ···················· |
| F | 7483 | hCoV-19/Wales/PHWC-169453/2020\|EPI_ISL_499445\|2020 | ···················· | ···················· | ···················· | ···················· | ···················· | ···················· |
| F | 7484 | hCoV-19/Wales/PHWC-16949F/2020\|EPI_ISL_499446\|2020 | ···················· | ···················· | ···················· | ···················· | ···················· | ···················· |
| F | 7485 | hCoV-19/Wales/PHWC-1694AE/2020\|EPI_ISL_499447\|2020 | ···················· | ···················· | ···················· | ···················· | ···················· | ···················· |
| F | 7486 | hCoV-19/Wales/PHWC-1694BD/2020\|EPI_ISL_499448\|2020 | ···················· | ···················· | ···················· | ···················· | ···················· | ···················· |
| F | 7487 | hCoV-19/Wales/PHWC-1694CC/2020\|EPI_ISL_499449\|2020 | ···················· | ···················· | ···················· | ···················· | ···················· | ···················· |

| | | | | | | | |
|---|---|---|---|---|---|---|---|
| F | 7488 hCoV-19/Wales/PHWC-1694EA/2020\|EPI_ISL_499450\|2020 | .................... | .................... | .................... | .................... | .................... | .................... |
| F | 7490 hCoV-19/Wales/PHWC-169505/2020\|EPI_ISL_499452\|2020 | .................... | .................... | .................... | .................... | .................... | .................... |
| F | 7491 hCoV-19/Wales/PHWC-169514/2020\|EPI_ISL_499453\|2020 | .................... | .................... | .................... | .................... | .................... | .................... |
| F | 7492 hCoV-19/Wales/PHWC-169523/2020\|EPI_ISL_499454\|2020 | .................... | .................... | .................... | .................... | .................... | .................... |
| F | 7494 hCoV-19/Wales/PHWC-16956F/2020\|EPI_ISL_499456\|2020 | .................... | .................... | .................... | .................... | .................... | .................... |
| F | 7495 hCoV-19/Wales/PHWC-16957E/2020\|EPI_ISL_499457\|2020 | .................... | .................... | .................... | .................... | .................... | .................... |
| F | 7496 hCoV-19/Wales/PHWC-16958D/2020\|EPI_ISL_499458\|2020 | .................... | .................... | .................... | .................... | .................... | .................... |
| F | 7497 hCoV-19/Wales/PHWC-1695AB/2020\|EPI_ISL_499459\|2020 | .................... | .................... | .................... | .................... | .................... | .................... |
| F | 7498 hCoV-19/England/LIVE-A586F/2020\|EPI_ISL_499461\|202 | .................... | .................... | .................... | .................... | .................... | .................... |
| F | 7502 hCoV-19/England/LIVE-A5F2E/2020\|EPI_ISL_499465\|202 | .................... | .................... | .................... | .................... | .................... | .................... |
| F | 7503 hCoV-19/England/LIVE-A62BB/2020\|EPI_ISL_499466\|202 | .................... | .................... | .................... | .................... | .................... | .................... |
| F | 7504 hCoV-19/England/LIVE-A67BC/2020\|EPI_ISL_499467\|202 | .................... | .................... | .................... | .................... | .................... | .................... |
| F | 7505 hCoV-19/Northern Ireland/NIRE-1069B3/2020\|EPI_ISL_ | .................... | .................... | .................... | .................... | .................... | .................... |
| F | 7506 hCoV-19/Northern Ireland/NIRE-106379/2020\|EPI_ISL_ | .................... | .................... | .................... | .................... | .................... | .................... |
| F | 7507 hCoV-19/Northern Ireland/NIRE-106C9C/2020\|EPI_ISL_ | .................... | .................... | .................... | .................... | .................... | .................... |
| F | 7508 hCoV-19/Northern Ireland/NIRE-106A83/2020\|EPI_ISL_ | .................... | .................... | .................... | .................... | .................... | .................... |
| F | 7509 hCoV-19/Northern Ireland/NIRE-106D99/2020\|EPI_ISL_ | .................... | .................... | .................... | .................... | .................... | .................... |
| F | 7510 hCoV-19/Northern Ireland/NIRE-1066E9/2020\|EPI_ISL_ | .................... | .................... | .................... | .................... | .................... | .................... |
| F | 7511 hCoV-19/Northern Ireland/NIRE-1064D0/2020\|EPI_ISL_ | .................... | .................... | .................... | .................... | .................... | .................... |
| F | 7512 hCoV-19/Northern Ireland/NIRE-10633D/2020\|EPI_ISL_ | .................... | .................... | .................... | .................... | .................... | .................... |
| F | 7513 hCoV-19/Northern Ireland/NIRE-1067F5/2020\|EPI_ISL_ | .................... | .................... | .................... | .................... | .................... | .................... |
| F | 7514 hCoV-19/Northern Ireland/NIRE-1069E0/2020\|EPI_ISL_ | .................... | .................... | .................... | .................... | .................... | .................... |
| F | 7515 hCoV-19/Northern Ireland/NIRE-1063B5/2020\|EPI_ISL_ | .................... | .................... | .................... | .................... | .................... | .................... |
| F | 7516 hCoV-19/Northern Ireland/NIRE-106DA8/2020\|EPI_ISL_ | .................... | .................... | .................... | .................... | .................... | .................... |
| F | 7517 hCoV-19/Northern Ireland/NIRE-1066DA/2020\|EPI_ISL_ | .................... | .................... | .................... | .................... | .................... | .................... |
| F | 7518 hCoV-19/Northern Ireland/NIRE-106BF9/2020\|EPI_ISL_ | .................... | .................... | .................... | .................... | .................... | .................... |
| F | 7519 hCoV-19/Northern Ireland/NIRE-106A47/2020\|EPI_ISL_ | .................... | .................... | .................... | .................... | .................... | .................... |
| F | 7520 hCoV-19/Northern Ireland/NIRE-106A92/2020\|EPI_ISL_ | .................... | .................... | .................... | .................... | .................... | .................... |
| F | 7521 hCoV-19/Northern Ireland/NIRE-106CC9/2020\|EPI_ISL_ | .................... | .................... | .................... | .................... | .................... | .................... |
| F | 7522 hCoV-19/Northern Ireland/NIRE-106A65/2020\|EPI_ISL_ | .................... | .................... | .................... | .................... | .................... | .................... |
| F | 7523 hCoV-19/Northern Ireland/NIRE-10677D/2020\|EPI_ISL_ | .................... | .................... | .................... | .................... | .................... | .................... |
| F | 7524 hCoV-19/Northern Ireland/NIRE-106DC6/2020\|EPI_ISL_ | .................... | .................... | .................... | .................... | .................... | .................... |
| F | 7525 hCoV-19/Northern Ireland/NIRE-106546/2020\|EPI_ISL_ | .................... | .................... | .................... | .................... | .................... | .................... |
| F | 7526 hCoV-19/Northern Ireland/NIRE-106D20/2020\|EPI_ISL_ | .................... | .................... | .................... | .................... | .................... | .................... |
| F | 7527 hCoV-19/Northern Ireland/NIRE-106A38/2020\|EPI_ISL_ | .................... | .................... | .................... | .................... | .................... | .................... |
| F | 7528 hCoV-19/Northern Ireland/NIRE-106652/2020\|EPI_ISL_ | .................... | .................... | .................... | .................... | .................... | .................... |
| F | 7529 hCoV-19/Northern Ireland/NIRE-106CAB/2020\|EPI_ISL_ | .................... | .................... | .................... | .................... | .................... | .................... |
| F | 7530 hCoV-19/Northern Ireland/NIRE-1064B2/2020\|EPI_ISL_ | .................... | .................... | .................... | .................... | .................... | .................... |
| F | 7531 hCoV-19/Northern Ireland/NIRE-106977/2020\|EPI_ISL_ | .................... | .................... | .................... | .................... | .................... | .................... |
| F | 7532 hCoV-19/Northern Ireland/NIRE-10687A/2020\|EPI_ISL_ | .................... | .................... | .................... | .................... | .................... | .................... |
| F | 7533 hCoV-19/Northern Ireland/NIRE-1064FE/2020\|EPI_ISL_ | .................... | .................... | .................... | .................... | .................... | .................... |
| F | 7534 hCoV-19/Northern Ireland/NIRE-106F1B/2020\|EPI_ISL_ | .................... | .................... | .................... | .................... | .................... | .................... |
| F | 7535 hCoV-19/Northern Ireland/NIRE-1069A4/2020\|EPI_ISL_ | .................... | .................... | .................... | .................... | .................... | .................... |
| F | 7536 hCoV-19/Northern Ireland/NIRE-106C05/2020\|EPI_ISL_ | .................... | .................... | .................... | .................... | .................... | .................... |
| F | 7537 hCoV-19/Northern Ireland/NIRE-106C32/2020\|EPI_ISL_ | .................... | .................... | .................... | .................... | .................... | .................... |
| F | 7538 hCoV-19/USA/NM-UNM-00348/2020\|EPI_ISL_508002\|2020- | .................... | .................... | .................... | .................... | .................... | .................... |
| F | 7539 hCoV-19/South Africa/KRISP-K000659/2020\|EPI_ISL_50 | .................... | .................... | .................... | .................... | .................... | .................... |
| F | 7540 hCoV-19/South Africa/KRISP-K001035/2020\|EPI_ISL_50 | .................... | .................... | .................... | .................... | .................... | .................... |
| F | 7541 hCoV-19/South Africa/KRISP-K001037/2020\|EPI_ISL_50 | .................... | .................... | .................... | .................... | .................... | .................... |
| F | 7542 hCoV-19/South Africa/KRISP-K001038/2020\|EPI_ISL_50 | .................... | .................... | .................... | .................... | .................... | .................... |
| F | 7543 hCoV-19/South Africa/KRISP-K001041/2020\|EPI_ISL_50 | .................... | .................... | .................... | .................... | .................... | .................... |
| F | 7544 hCoV-19/South Africa/KRISP-K001043/2020\|EPI_ISL_50 | .................... | .................... | .................... | .................... | .................... | .................... |
| F | 7545 hCoV-19/South Africa/KRISP-K001044/2020\|EPI_ISL_50 | .................... | .................... | .................... | .................... | .................... | .................... |
| F | 7546 hCoV-19/South Africa/KRISP-K001045/2020\|EPI_ISL_50 | .................... | .................... | .................... | .................... | .................... | .................... |
| F | 7547 hCoV-19/South Africa/KRISP-K001049/2020\|EPI_ISL_50 | .................... | .................... | .................... | .................... | .................... | .................... |
| F | 7548 hCoV-19/South Africa/KRISP-K001051/2020\|EPI_ISL_50 | .................... | .................... | .................... | .................... | .................... | .................... |
| F | 7549 hCoV-19/South Africa/KRISP-K001052/2020\|EPI_ISL_50 | .................... | .................... | .................... | .................... | .................... | .................... |
| F | 7550 hCoV-19/South Africa/KRISP-K001053/2020\|EPI_ISL_50 | .................... | .................... | .................... | .................... | .................... | .................... |
| F | 7551 hCoV-19/South Africa/KRISP-K001054/2020\|EPI_ISL_50 | .................... | .................... | .................... | .................... | .................... | .................... |
| F | 7552 hCoV-19/South Africa/KRISP-K001055/2020\|EPI_ISL_50 | .................... | .................... | .................... | .................... | .................... | .................... |
| F | 7553 hCoV-19/South Africa/KRISP-K001057/2020\|EPI_ISL_50 | .................... | .................... | .................... | .................... | .................... | .................... |
| F | 7554 hCoV-19/South Africa/KRISP-K001058/2020\|EPI_ISL_50 | .................... | .................... | .................... | .................... | .................... | .................... |
| F | 7555 hCoV-19/South Africa/KRISP-K001059/2020\|EPI_ISL_50 | .................... | .................... | .................... | .................... | .................... | .................... |
| F | 7556 hCoV-19/South Africa/KRISP-K001061/2020\|EPI_ISL_50 | .................... | .................... | .................... | .................... | .................... | .................... |
| F | 7557 hCoV-19/South Africa/KRISP-K001062/2020\|EPI_ISL_50 | .................... | .................... | .................... | .................... | .................... | .................... |
| F | 7558 hCoV-19/South Africa/KRISP-K001063/2020\|EPI_ISL_50 | .................... | .................... | .................... | .................... | .................... | .................... |
| F | 7559 hCoV-19/South Africa/KRISP-K001064/2020\|EPI_ISL_50 | .................... | .................... | .................... | .................... | .................... | .................... |
| F | 7560 hCoV-19/South Africa/KRISP-K001065/2020\|EPI_ISL_50 | .................... | .................... | .................... | .................... | .................... | .................... |
| F | 7561 hCoV-19/South Africa/KRISP-K001066/2020\|EPI_ISL_50 | .................... | .................... | .................... | .................... | .................... | .................... |
| F | 7562 hCoV-19/South Africa/KRISP-K001067/2020\|EPI_ISL_50 | .................... | .................... | .................... | .................... | .................... | .................... |
| F | 7563 hCoV-19/South Africa/KRISP-K001068/2020\|EPI_ISL_50 | .................... | .................... | .................... | .................... | .................... | .................... |
| F | 7564 hCoV-19/South Africa/KRISP-K001071/2020\|EPI_ISL_50 | .................... | .................... | .................... | .................... | .................... | .................... |
| F | 7565 hCoV-19/South Africa/KRISP-K001072/2020\|EPI_ISL_50 | .................... | .................... | .................... | .................... | .................... | .................... |
| F | 7566 hCoV-19/South Africa/KRISP-K001073/2020\|EPI_ISL_50 | .................... | .................... | .................... | .................... | .................... | .................... |
| F | 7567 hCoV-19/South Africa/KRISP-K001074/2020\|EPI_ISL_50 | .................... | .................... | .................... | .................... | .................... | .................... |
| F | 7568 hCoV-19/South Africa/KRISP-K001076/2020\|EPI_ISL_50 | .................... | .................... | .................... | .................... | .................... | .................... |
| F | 7569 hCoV-19/South Africa/KRISP-K001077/2020\|EPI_ISL_50 | .................... | .................... | .................... | .................... | .................... | .................... |
| F | 7570 hCoV-19/South Africa/KRISP-K001078/2020\|EPI_ISL_50 | .................... | .................... | .................... | .................... | .................... | .................... |
| F | 7571 hCoV-19/South Africa/KRISP-K001079/2020\|EPI_ISL_50 | .................... | .................... | .................... | .................... | .................... | .................... |
| F | 7572 hCoV-19/South Africa/KRISP-K001080/2020\|EPI_ISL_50 | .................... | .................... | .................... | .................... | .................... | .................... |

| | | | | | | | | |
|---|---|---|---|---|---|---|---|---|
| F | 7573 | hCoV-19/South Africa/KRISP-K001082/2020\|EPI_ISL_50 | ···················· | ···················· | ···················· | ···················· | ···················· | ···················· |
| F | 7574 | hCoV-19/South Africa/KRISP-K001083/2020\|EPI_ISL_50 | ···················· | ···················· | ···················· | ···················· | ···················· | ···················· |
| F | 7575 | hCoV-19/South Africa/KRISP-K001084/2020\|EPI_ISL_50 | ···················· | ···················· | ···················· | ···················· | ···················· | ···················· |
| F | 7576 | hCoV-19/South Africa/KRISP-K001085/2020\|EPI_ISL_50 | ···················· | ···················· | ···················· | ···················· | ···················· | ···················· |
| F | 7577 | hCoV-19/South Africa/KRISP-K001087/2020\|EPI_ISL_50 | ···················· | ···················· | ···················· | ···················· | ···················· | ···················· |
| F | 7578 | hCoV-19/Spain/GA-IBV-004062/2020\|EPI_ISL_510300\|20 | ···················· | ···················· | ···················· | ···················· | ···················· | ···················· |
| F | 7579 | hCoV-19/Spain/GA-IBV-004063/2020\|EPI_ISL_510301\|20 | ···················· | ···················· | ···················· | ···················· | ···················· | ···················· |
| F | 7580 | hCoV-19/Spain/GA-IBV-004064/2020\|EPI_ISL_510302\|20 | ···················· | ···················· | ···················· | ···················· | ···················· | ···················· |
| F | 7581 | hCoV-19/Spain/GA-IBV-004065/2020\|EPI_ISL_510303\|20 | ···················· | ···················· | ···················· | ···················· | ···················· | ···················· |
| F | 7593 | hCoV-19/Spain/EX-IBV-005008/2020\|EPI_ISL_510317\|20 | ···················· | ···················· | ···················· | ···················· | ···················· | ···················· |
| F | 7594 | hCoV-19/Spain/EX-IBV-005011/2020\|EPI_ISL_510320\|20 | ···················· | ···················· | ···················· | ···················· | ···················· | ···················· |
| F | 7597 | hCoV-19/Spain/EX-IBV-005014/2020\|EPI_ISL_510323\|20 | ···················· | ···················· | ···················· | ···················· | ···················· | ···················· |
| F | 7599 | hCoV-19/Portugal/PT1065/2020\|EPI_ISL_511612\|2020-0 | ···················· | ···················· | ···················· | ···················· | ···················· | ···················· |
| F | 7600 | hCoV-19/Portugal/PT1066/2020\|EPI_ISL_511613\|2020-0 | ···················· | ···················· | ···················· | ···················· | ···················· | ···················· |
| F | 7601 | hCoV-19/Portugal/PT1067/2020\|EPI_ISL_511614\|2020-0 | ···················· | ···················· | ···················· | ···················· | ···················· | ···················· |
| F | 7602 | hCoV-19/Portugal/PT1068/2020\|EPI_ISL_511615\|2020-0 | ···················· | ···················· | ···················· | ···················· | ···················· | ···················· |
| F | 7603 | hCoV-19/Portugal/PT1069/2020\|EPI_ISL_511616\|2020-0 | ···················· | ···················· | ···················· | ···················· | ···················· | ···················· |
| F | 7604 | hCoV-19/Portugal/PT1070/2020\|EPI_ISL_511617\|2020-0 | ···················· | ···················· | ···················· | ···················· | ···················· | ···················· |
| F | 7605 | hCoV-19/Portugal/PT1072/2020\|EPI_ISL_511619\|2020-0 | ···················· | ···················· | ···················· | ···················· | ···················· | ···················· |
| F | 7606 | hCoV-19/Portugal/PT1073/2020\|EPI_ISL_511620\|2020-0 | ···················· | ···················· | ···················· | ···················· | ···················· | ···················· |
| F | 7607 | hCoV-19/Portugal/PT1074/2020\|EPI_ISL_511621\|2020-0 | ···················· | ···················· | ···················· | ···················· | ···················· | ···················· |
| F | 7608 | hCoV-19/Portugal/PT1075/2020\|EPI_ISL_511622\|2020-0 | ···················· | ···················· | ···················· | ···················· | ···················· | ···················· |
| F | 7609 | hCoV-19/Portugal/PT1076/2020\|EPI_ISL_511623\|2020-0 | ···················· | ···················· | ···················· | ···················· | ···················· | ···················· |
| F | 7610 | hCoV-19/Portugal/PT1077/2020\|EPI_ISL_511624\|2020-0 | ···················· | ···················· | ···················· | ···················· | ···················· | ···················· |
| F | 7611 | hCoV-19/Portugal/PT1078/2020\|EPI_ISL_511625\|2020-0 | ···················· | ···················· | ···················· | ···················· | ···················· | ···················· |
| F | 7612 | hCoV-19/Portugal/PT1079/2020\|EPI_ISL_511626\|2020-0 | ···················· | ···················· | ···················· | ···················· | ···················· | ···················· |
| F | 7613 | hCoV-19/Portugal/PT1080/2020\|EPI_ISL_511627\|2020-0 | ···················· | ···················· | ···················· | ···················· | ···················· | ···················· |
| F | 7614 | hCoV-19/Portugal/PT1081/2020\|EPI_ISL_511628\|2020-0 | ···················· | ···················· | ···················· | ···················· | ···················· | ···················· |
| F | 7615 | hCoV-19/Portugal/PT1082/2020\|EPI_ISL_511629\|2020-0 | ···················· | ···················· | ···················· | ···················· | ···················· | ···················· |
| F | 7616 | hCoV-19/Portugal/PT1083/2020\|EPI_ISL_511630\|2020-0 | ···················· | ···················· | ···················· | ···················· | ···················· | ···················· |
| F | 7617 | hCoV-19/Portugal/PT1084/2020\|EPI_ISL_511631\|2020-0 | ···················· | ···················· | ···················· | ···················· | ···················· | ···················· |
| F | 7618 | hCoV-19/Portugal/PT1085/2020\|EPI_ISL_511632\|2020-0 | ···················· | ···················· | ···················· | ···················· | ···················· | ···················· |
| F | 7619 | hCoV-19/Portugal/PT1086/2020\|EPI_ISL_511633\|2020-0 | ···················· | ···················· | ···················· | ···················· | ···················· | ···················· |
| F | 7620 | hCoV-19/Portugal/PT1087/2020\|EPI_ISL_511634\|2020-0 | ···················· | ···················· | ···················· | ···················· | ···················· | ···················· |
| F | 7621 | hCoV-19/Portugal/PT1088/2020\|EPI_ISL_511635\|2020-0 | ···················· | ···················· | ···················· | ···················· | ···················· | ···················· |
| F | 7622 | hCoV-19/Portugal/PT1089/2020\|EPI_ISL_511636\|2020-0 | ···················· | ···················· | ···················· | ···················· | ···················· | ···················· |
| F | 7623 | hCoV-19/Portugal/PT1090/2020\|EPI_ISL_511637\|2020-0 | ···················· | ···················· | ···················· | ···················· | ···················· | ···················· |
| F | 7624 | hCoV-19/Portugal/PT1091/2020\|EPI_ISL_511638\|2020-0 | ···················· | ···················· | ···················· | ···················· | ···················· | ···················· |
| F | 7625 | hCoV-19/Portugal/PT1092/2020\|EPI_ISL_511639\|2020-0 | ···················· | ···················· | ···················· | ···················· | ···················· | ···················· |
| F | 7626 | hCoV-19/Portugal/PT1093/2020\|EPI_ISL_511640\|2020-0 | ···················· | ···················· | ···················· | ···················· | ···················· | ···················· |
| F | 7627 | hCoV-19/Portugal/PT1095/2020\|EPI_ISL_511642\|2020-0 | ···················· | ···················· | ···················· | ···················· | ···················· | ···················· |
| F | 7628 | hCoV-19/Portugal/PT1096/2020\|EPI_ISL_511643\|2020-0 | ···················· | ···················· | ···················· | ···················· | ···················· | ···················· |
| F | 7629 | hCoV-19/Portugal/PT1098/2020\|EPI_ISL_511645\|2020-0 | ···················· | ···················· | ···················· | ···················· | ···················· | ···················· |
| F | 7630 | hCoV-19/Portugal/PT1099/2020\|EPI_ISL_511646\|2020-0 | ···················· | ···················· | ···················· | ···················· | ···················· | ···················· |
| F | 7631 | hCoV-19/Portugal/PT1100/2020\|EPI_ISL_511647\|2020-0 | ···················· | ···················· | ···················· | ···················· | ···················· | ···················· |
| F | 7632 | hCoV-19/Portugal/PT1101/2020\|EPI_ISL_511648\|2020-0 | ···················· | ····A··············· | ···················· | ···················· | ···················· | ···················· |
| F | 7633 | hCoV-19/Portugal/PT1103/2020\|EPI_ISL_511650\|2020-0 | ···················· | ···················· | ···················· | ···················· | ···················· | ···················· |
| F | 7634 | hCoV-19/Portugal/PT1104/2020\|EPI_ISL_511651\|2020-0 | ···················· | ···················· | ···················· | ···················· | ···················· | ···················· |
| F | 7635 | hCoV-19/Portugal/PT1105/2020\|EPI_ISL_511652\|2020-0 | ···················· | ···················· | ···················· | ···················· | ···················· | ···················· |
| F | 7636 | hCoV-19/Portugal/PT1106/2020\|EPI_ISL_511653\|2020-0 | ···················· | ···················· | ···················· | ···················· | ···················· | ···················· |
| F | 7637 | hCoV-19/Portugal/PT1107/2020\|EPI_ISL_511654\|2020-0 | ···················· | ···················· | ···················· | ···················· | ···················· | ···················· |
| F | 7638 | hCoV-19/Portugal/PT1108/2020\|EPI_ISL_511655\|2020-0 | ···················· | ···················· | ···················· | ···················· | ···················· | ···················· |
| F | 7639 | hCoV-19/SaudiArabia/KAUST-JEDDAH506/2020\|EPI_ISL_5 | ···················· | ···················· | ···················· | ···················· | ···················· | ···················· |
| F | 7640 | hCoV-19/SaudiArabia/KAUST-JEDDAH525/2020\|EPI_ISL_5 | ···················· | ···················· | ···················· | ···················· | ···················· | ···················· |
| F | 7641 | hCoV-19/SaudiArabia/KAUST-JEDDAH528/2020\|EPI_ISL_5 | ···················· | ···················· | ···················· | ···················· | ···················· | ···················· |
| F | 7642 | hCoV-19/SaudiArabia/KAUST-JEDDAH529/2020\|EPI_ISL_5 | ···················· | ···················· | ···················· | ···················· | ···················· | ···················· |
| F | 7643 | hCoV-19/SaudiArabia/KAUST-JEDDAH531/2020\|EPI_ISL_5 | ···················· | ···················· | ···················· | ···················· | ···················· | ···················· |
| F | 7644 | hCoV-19/SaudiArabia/KAUST-JEDDAH534/2020\|EPI_ISL_5 | ···················· | ···················· | ···················· | ···················· | ···················· | ···················· |
| F | 7645 | hCoV-19/SaudiArabia/KAUST-JEDDAH535/2020\|EPI_ISL_5 | ···················· | ···················· | ···················· | ···················· | ···················· | ···················· |
| F | 7646 | hCoV-19/SaudiArabia/KAUST-JEDDAH536/2020\|EPI_ISL_5 | ···················· | ···················· | ···················· | ···················· | ···················· | ···················· |
| F | 7647 | hCoV-19/SaudiArabia/KAUST-JEDDAH538/2020\|EPI_ISL_5 | ···················· | ···················· | ···················· | ···················· | ···················· | ···················· |
| F | 7648 | hCoV-19/SaudiArabia/KAUST-JEDDAH539/2020\|EPI_ISL_5 | ···················· | ·A·················· | ···················· | ···················· | ···················· | ···A················ |
| F | 7649 | hCoV-19/SaudiArabia/KAUST-JEDDAH542/2020\|EPI_ISL_5 | ···················· | ···················· | ···················· | ···················· | ···················· | ···················· |
| F | 7650 | hCoV-19/SaudiArabia/KAUST-JEDDAH544/2020\|EPI_ISL_5 | ···················· | ···················· | ···················· | ···················· | ···················· | ···················· |
| F | 7651 | hCoV-19/SaudiArabia/KAUST-JEDDAH545/2020\|EPI_ISL_5 | ···················· | ···················· | ···················· | ···················· | ···················· | ···················· |
| F | 7652 | hCoV-19/SaudiArabia/KAUST-JEDDAH546/2020\|EPI_ISL_5 | ···················· | ···················· | ···················· | ···················· | ···················· | ···················· |
| F | 7653 | hCoV-19/SaudiArabia/KAUST-JEDDAH547/2020\|EPI_ISL_5 | ···················· | ···················· | ···················· | ···················· | ···················· | ···················· |
| F | 7654 | hCoV-19/SaudiArabia/KAUST-JEDDAH558/2020\|EPI_ISL_5 | ···················· | ···················· | ···················· | ···················· | ···················· | ···················· |
| F | 7655 | hCoV-19/SaudiArabia/KAUST-JEDDAH566/2020\|EPI_ISL_5 | ···················· | ···················· | ···················· | ···················· | ···················· | ···················· |
| F | 7656 | hCoV-19/SaudiArabia/KAUST-JEDDAH569/2020\|EPI_ISL_5 | ···················· | ···················· | ···················· | ···················· | ···················· | ···················· |
| F | 7657 | hCoV-19/Saudi Arabia/KAUST-JEDDAH572/2020\|EPI_ISL_ | ···················· | ···················· | ···················· | ···················· | ···················· | ···················· |
| F | 7658 | hCoV-19/SaudiArabia/KAUST-JEDDAH573/2020\|EPI_ISL_5 | ···················· | ···················· | ···················· | ···················· | ···················· | ···················· |
| F | 7659 | hCoV-19/Saudi Arabia/KAUST-JEDDAH576/2020\|EPI_ISL_ | ···················· | ···················· | ···················· | ···················· | ···················· | ···················· |
| F | 7660 | hCoV-19/SaudiArabia/KAUST-JEDDAH577/2020\|EPI_ISL_5 | ···················· | ···················· | ···················· | ···················· | ···················· | ···················· |
| F | 7661 | hCoV-19/SaudiArabia/KAUST-JEDDAH578/2020\|EPI_ISL_5 | ···················· | ···················· | ···················· | ···················· | ···················· | ···················· |
| F | 7662 | hCoV-19/SaudiArabia/KAUST-JEDDAH581/2020\|EPI_ISL_5 | ···················· | ···················· | ···················· | ···················· | ···················· | ···················· |
| F | 7663 | hCoV-19/SaudiArabia/KAUST-JEDDAH582/2020\|EPI_ISL_5 | ···················· | ···················· | ···················· | ···················· | ···················· | ···················· |
| F | 7664 | hCoV-19/SaudiArabia/KAUST-JEDDAH583/2020\|EPI_ISL_5 | ···················· | ···················· | ···················· | ···················· | ···················· | ···················· |
| F | 7665 | hCoV-19/SaudiArabia/KAUST-JEDDAH586/2020\|EPI_ISL_5 | ···················· | ···················· | ···················· | ···················· | ···················· | ···················· |
| F | 7666 | hCoV-19/SaudiArabia/KAUST-JEDDAH587/2020\|EPI_ISL_5 | ···················· | ···················· | ···················· | ···················· | ···················· | ···················· |

| | | | | | | | | |
|---|---|---|---|---|---|---|---|---|
| F | 7667 | hCoV-19/SaudiArabia/KAUST-JEDDAH589/2020\|EPI_ISL_5 | ···················· | ···················· | ···················· | ···················· | ···················· | ···················· |
| F | 7668 | hCoV-19/SaudiArabia/KAUST-JEDDAH590/2020\|EPI_ISL_5 | ···················· | ···················· | ···················· | ···················· | ···················· | ···················· |
| F | 7669 | hCoV-19/SaudiArabia/KAUST-JEDDAH591/2020\|EPI_ISL_5 | ···················· | ···················· | ···················· | ···················· | ···················· | ···················· |
| F | 7670 | hCoV-19/SaudiArabia/KAUST-JEDDAH592/2020\|EPI_ISL_5 | ···················· | ···················· | ···················· | ···················· | ···················· | ···················· |
| F | 7671 | hCoV-19/SaudiArabia/KAUST-JEDDAH594/2020\|EPI_ISL_5 | ···················· | ···················· | ···················· | ···················· | ···················· | ···················· |
| F | 7672 | hCoV-19/SaudiArabia/KAUST-JEDDAH595/2020\|EPI_ISL_5 | ···················· | ···················· | ···················· | ···················· | ···················· | ···················· |
| F | 7673 | hCoV-19/SaudiArabia/KAUST-JEDDAH613/2020\|EPI_ISL_5 | ···················· | ···················· | ···················· | ···················· | ···················· | ···················· |
| F | 7674 | hCoV-19/USA/WI-UW-1074/2020\|EPI_ISL_516500\|2020-07 | ···················· | ···················· | ···················· | ···················· | ···················· | ···················· |
| F | 7675 | hCoV-19/USA/IL-UW-1075/2020\|EPI_ISL_516501\|2020-07 | ···················· | ···················· | ···················· | ···················· | ···················· | ···················· |
| F | 7676 | hCoV-19/USA/WI-UW-1076/2020\|EPI_ISL_516502\|2020-07 | ···················· | ···················· | ···················· | ···················· | ···················· | ···················· |
| F | 7677 | hCoV-19/USA/WI-UW-1077/2020\|EPI_ISL_516503\|2020-07 | ···················· | ···················· | ···················· | ···················· | ···················· | ···················· |
| F | 7678 | hCoV-19/USA/WI-UW-1078/2020\|EPI_ISL_516504\|2020-07 | ···················· | ·A·················· | ···················· | ···················· | ···················· | ···················· |
| F | 7679 | hCoV-19/USA/WI-UW-1079/2020\|EPI_ISL_516505\|2020-07 | ···················· | ···················· | ···················· | ···················· | ···················· | ···················· |
| F | 7680 | hCoV-19/USA/WI-UW-1080/2020\|EPI_ISL_516506\|2020-08 | ···················· | ·A·················· | ···················· | ···················· | ···················· | ···················· |
| F | 7681 | hCoV-19/South Africa/KRISP-0489/2020\|EPI_ISL_49806 | ···················· | ···················· | ···················· | ···················· | ···················· | ···················· |
| F | 7682 | hCoV-19/South Africa/KRISP-0490/2020\|EPI_ISL_49806 | ···················· | ···················· | ···················· | ···················· | ···················· | ···················· |
| F | 7683 | hCoV-19/South Africa/KRISP-0491/2020\|EPI_ISL_49806 | ···················· | ···················· | ···················· | ···················· | ···················· | ···················· |
| F | 7684 | hCoV-19/South Africa/KRISP-0493/2020\|EPI_ISL_49806 | ···················· | ···················· | ···················· | ···················· | ···················· | ···················· |
| F | 7685 | hCoV-19/South Africa/KRISP-0494/2020\|EPI_ISL_49806 | ···················· | ···················· | ···················· | ···················· | ···················· | ···················· |
| F | 7686 | hCoV-19/South Africa/KRISP-0497/2020\|EPI_ISL_49806 | ···················· | ···················· | ···················· | ···················· | ···················· | ···················· |
| F | 7687 | hCoV-19/South Africa/KRISP-0498/2020\|EPI_ISL_49806 | ···················· | ···················· | ···················· | ···················· | ···················· | ···················· |
| F | 7688 | hCoV-19/South Africa/KRISP-0501/2020\|EPI_ISL_49806 | ···················· | ···················· | ···················· | ···················· | ···················· | ···················· |
| F | 7689 | hCoV-19/South Africa/KRISP-0502/2020\|EPI_ISL_49807 | ···················· | ···················· | ···················· | ···················· | ···················· | ···················· |
| F | 7690 | hCoV-19/South Africa/KRISP-0503/2020\|EPI_ISL_49807 | ···················· | ···················· | ···················· | ···················· | ···················· | ···················· |
| F | 7691 | hCoV-19/South Africa/KRISP-0526/2020\|EPI_ISL_49807 | ···················· | ···················· | ···················· | ···················· | ···················· | ···················· |
| F | 7692 | hCoV-19/South Africa/KRISP-0527/2020\|EPI_ISL_49807 | ···················· | ···················· | ···················· | ···················· | ···················· | ···················· |
| F | 7693 | hCoV-19/South Africa/KRISP-0529/2020\|EPI_ISL_49807 | ···················· | ···················· | ···················· | ···················· | ···················· | ···················· |
| F | 7694 | hCoV-19/South Africa/KRISP-0531/2020\|EPI_ISL_49807 | ···················· | ···················· | ···················· | ···················· | ···················· | ···················· |
| F | 7695 | hCoV-19/South Africa/KRISP-0534/2020\|EPI_ISL_49807 | ···················· | ···················· | ···················· | ···················· | ···················· | ···················· |
| F | 7696 | hCoV-19/South Africa/KRISP-0535/2020\|EPI_ISL_49807 | ···················· | ···················· | ···················· | ···················· | ···················· | ···················· |
| F | 7697 | hCoV-19/South Africa/KRISP-0536/2020\|EPI_ISL_49807 | ···················· | ···················· | ···················· | ···················· | ···················· | ···················· |
| F | 7698 | hCoV-19/South Africa/KRISP-0551/2020\|EPI_ISL_49809 | ···················· | ···················· | ···················· | ···················· | ···················· | ···················· |
| F | 7699 | hCoV-19/South Africa/KRISP-0552/2020\|EPI_ISL_49809 | ···················· | ···················· | ···················· | ···················· | ···················· | ···················· |
| F | 7700 | hCoV-19/South Africa/KRISP-0553/2020\|EPI_ISL_49809 | ···················· | ···················· | ···················· | ···················· | ···················· | ···················· |
| F | 7701 | hCoV-19/South Africa/KRISP-0554/2020\|EPI_ISL_49809 | ···················· | ···················· | ···················· | ···················· | ···················· | ···················· |
| F | 7702 | hCoV-19/South Africa/KRISP-0555/2020\|EPI_ISL_49809 | ···················· | ···················· | ···················· | ···················· | ···················· | ···················· |
| F | 7703 | hCoV-19/South Africa/KRISP-0556/2020\|EPI_ISL_49809 | ···················· | ···················· | ···················· | ···················· | ···················· | ···················· |
| F | 7704 | hCoV-19/South Africa/KRISP-0557/2020\|EPI_ISL_49809 | ···················· | ···················· | ···················· | ···················· | ···················· | ···················· |
| F | 7705 | hCoV-19/South Africa/KRISP-0558/2020\|EPI_ISL_49809 | ···················· | ···················· | ···················· | ···················· | ···················· | ···················· |
| F | 7706 | hCoV-19/South Africa/KRISP-0559/2020\|EPI_ISL_49809 | ···················· | ···················· | ···················· | ···················· | ···················· | ···················· |
| F | 7707 | hCoV-19/South Africa/KRISP-0560/2020\|EPI_ISL_49809 | ···················· | ···················· | ···················· | ···················· | ···················· | ···················· |
| F | 7708 | hCoV-19/Wales/PHWC-16909B/2020\|EPI_ISL_499390\|2020 | ···················· | ···················· | ···················· | ···················· | ···················· | ···················· |
| F | 7709 | hCoV-19/Wales/PHWC-1690D7/2020\|EPI_ISL_499393\|2020 | ···················· | ···················· | ···················· | ···················· | ···················· | ···················· |
| F | 7710 | hCoV-19/Wales/PHWC-169110/2020\|EPI_ISL_499395\|2020 | ···················· | ···················· | ···················· | ···················· | ···················· | ···················· |
| F | 7711 | hCoV-19/Wales/PHWC-16912F/2020\|EPI_ISL_499396\|2020 | ···················· | ···················· | ···················· | ···················· | ···················· | ···················· |
| F | 7712 | hCoV-19/Wales/PHWC-1692C2/2020\|EPI_ISL_499398\|2020 | ···················· | ···················· | ···················· | ···················· | ···················· | ···················· |
| F | 7713 | hCoV-19/Wales/PHWC-169259/2020\|EPI_ISL_499399\|2020 | ···················· | ···················· | ···················· | ···················· | ···················· | ···················· |
| F | 7714 | hCoV-19/Wales/PHWC-16914D/2020\|EPI_ISL_499415\|2020 | ···················· | ···················· | ···················· | ···················· | ···················· | ···················· |
| F | 7715 | hCoV-19/Wales/PHWC-16915C/2020\|EPI_ISL_499416\|2020 | ···················· | ···················· | ···················· | ···················· | ···················· | ···················· |
| F | 7716 | hCoV-19/Wales/PHWC-1691D4/2020\|EPI_ISL_499418\|2020 | ···················· | ···················· | ···················· | ···················· | ···················· | ···················· |
| F | 7717 | hCoV-19/Wales/PHWC-1691E3/2020\|EPI_ISL_499419\|2020 | ···················· | ···················· | ···················· | ···················· | ···················· | ···················· |
| F | 7718 | hCoV-19/Wales/PHWC-1691F2/2020\|EPI_ISL_499420\|2020 | ···················· | ···················· | ···················· | ···················· | ···················· | ···················· |
| F | 7719 | hCoV-19/Wales/PHWC-16920E/2020\|EPI_ISL_499421\|2020 | ···················· | ···················· | ···················· | ···················· | ···················· | ···················· |
| F | 7720 | hCoV-19/Wales/PHWC-16921D/2020\|EPI_ISL_499422\|2020 | ···················· | ···················· | ···················· | ···················· | ···················· | ···················· |
| F | 7721 | hCoV-19/Wales/PHWC-16924A/2020\|EPI_ISL_499423\|2020 | ···················· | ···················· | ···················· | ···················· | ···················· | ···················· |
| F | 7722 | hCoV-19/USA/NM-UNM-00347/2020\|EPI_ISL_508057\|2020- | ···················· | ···················· | ···················· | ···················· | ···················· | ···················· |
| F | 7723 | hCoV-19/USA/NM-UNM-00324/2020\|EPI_ISL_508063\|2020- | ···················· | ···················· | ···················· | ···················· | ···················· | ···················· |
| F | 7724 | hCoV-19/USA/NM-UNM-00320/2020\|EPI_ISL_508066\|2020- | ···················· | ···················· | ···················· | ···················· | ···················· | ···················· |
| F | 7725 | hCoV-19/USA/NM-UNM-00403/2020\|EPI_ISL_508091\|2020- | ···················· | ···················· | ···················· | ···················· | ···················· | ···················· |
| F | 7726 | hCoV-19/USA/NM-UNM-00341/2020\|EPI_ISL_508093\|2020- | ···················· | ···················· | ···················· | ···················· | ···················· | ···················· |
| F | 7727 | hCoV-19/USA/NM-UNM-00317/2020\|EPI_ISL_508097\|2020- | ···················· | ···················· | ···················· | ···················· | ···················· | ···················· |
| F | 7728 | hCoV-19/USA/NM-UNM-00325/2020\|EPI_ISL_508099\|2020- | ···················· | ···················· | ···················· | ···················· | ···················· | ···················· |
| F | 7729 | hCoV-19/Singapore/770/2020\|EPI_ISL_509380\|2020-07- | ···················· | ···················· | ···················· | ···················· | ···················· | ···················· |
| F | 7730 | hCoV-19/Singapore/771/2020\|EPI_ISL_509381\|2020-07- | ···················· | ···················· | ···················· | ···················· | ···················· | ···················· |
| F | 7731 | hCoV-19/Singapore/772/2020\|EPI_ISL_509382\|2020-07- | ···················· | ···················· | ···················· | ···················· | ···················· | ···················· |
| F | 7732 | hCoV-19/Singapore/773/2020\|EPI_ISL_509383\|2020-07- | ···················· | ···················· | ···················· | ···················· | ···················· | ···················· |
| F | 7733 | hCoV-19/Singapore/774/2020\|EPI_ISL_509384\|2020-07- | ···················· | ···················· | ···················· | ···················· | ···················· | ···················· |
| F | 7734 | hCoV-19/Singapore/775/2020\|EPI_ISL_509385\|2020-07- | ···················· | ·A·················· | ···················· | ···················· | ···················· | ···················· |
| F | 7735 | hCoV-19/Singapore/776/2020\|EPI_ISL_509386\|2020-07- | ···················· | ···················· | ···················· | ···················· | ···················· | ···················· |
| F | 7736 | hCoV-19/Singapore/777/2020\|EPI_ISL_509387\|2020-07- | ···················· | ·A·················· | ···················· | ···················· | ···················· | ···A················ |
| F | 7737 | hCoV-19/Singapore/778/2020\|EPI_ISL_509388\|2020-07- | ···················· | ···················· | ···················· | ···················· | ···················· | ···················· |
| F | 7738 | hCoV-19/Singapore/779/2020\|EPI_ISL_509389\|2020-07- | ···················· | ···················· | ···················· | ···················· | ···················· | ···················· |
| F | 7739 | hCoV-19/Singapore/780/2020\|EPI_ISL_509390\|2020-07- | ···················· | ···················· | ···················· | ···················· | ···················· | ···················· |
| F | 7740 | hCoV-19/Singapore/781/2020\|EPI_ISL_509391\|2020-06- | ···················· | ···················· | ···················· | ···················· | ···················· | ···················· |
| F | 7741 | hCoV-19/Singapore/782/2020\|EPI_ISL_509392\|2020-06- | ···················· | ···················· | ···················· | ···················· | ···················· | ···················· |
| F | 7742 | hCoV-19/Singapore/783/2020\|EPI_ISL_509393\|2020-06- | ···················· | ···················· | ···················· | ···················· | ···················· | ···················· |
| F | 7743 | hCoV-19/Singapore/784/2020\|EPI_ISL_509394\|2020-06- | ···················· | ···················· | ···················· | ···················· | ···················· | ···················· |
| F | 7744 | hCoV-19/Turkey/ACUTG-5/2020\|EPI_ISL_509416\|2020-04 | ···················· | ···················· | ···················· | ···················· | ···················· | ···················· |
| F | 7754 | hCoV-19/Spain/AR-IBV-005112/2020\|EPI_ISL_510382\|20 | ···················· | ···················· | ···················· | ···················· | ···················· | ···················· |
| F | 7760 | hCoV-19/Spain/AR-IBV-005132/2020\|EPI_ISL_510390\|20 | ···················· | ···················· | ···················· | ···················· | ···················· | ···················· |

| | | | | | | | | |
|---|---|---|---|---|---|---|---|---|
| F | 7762 | hCoV-19/Spain/PV-IBV-004273/2020\|EPI_ISL_510392\|20 | .................... | .................... | ..................... | .................... | .................... | .................... |
| F | 7763 | hCoV-19/Spain/PV-IBV-004274/2020\|EPI_ISL_510393\|20 | .................... | .................... | ..................... | .................... | .................... | .................... |
| F | 7772 | hCoV-19/Portugal/PT1155/2020\|EPI_ISL_511702\|2020-0 | .................... | .................... | ..................... | .................... | .................... | .................... |
| F | 7773 | hCoV-19/Portugal/PT1159/2020\|EPI_ISL_511706\|2020-0 | .................... | .................... | ..................... | .................... | .................... | .................... |
| F | 7774 | hCoV-19/Portugal/PT1161/2020\|EPI_ISL_511708\|2020-0 | .................... | .................... | ..................... | .................... | .................... | .................... |
| F | 7775 | hCoV-19/Portugal/PT1162/2020\|EPI_ISL_511709\|2020-0 | .................... | .A.................. | ..................... | .................... | .................... | .................... |
| F | 7776 | hCoV-19/Portugal/PT1163/2020\|EPI_ISL_511710\|2020-0 | .................... | .................... | ..................... | .................... | .................... | .................... |
| F | 7777 | hCoV-19/Portugal/PT1165/2020\|EPI_ISL_511711\|2020-0 | .................... | .................... | ..................... | .................... | .................... | .................... |
| F | 7778 | hCoV-19/Portugal/PT1167/2020\|EPI_ISL_511713\|2020-0 | .................... | .................... | ..................... | .................... | .................... | .................... |
| F | 7779 | hCoV-19/Portugal/PT1168/2020\|EPI_ISL_511714\|2020-0 | .................... | .................... | ..................... | .................... | .................... | .................... |
| F | 7780 | hCoV-19/Portugal/PT1169/2020\|EPI_ISL_511715\|2020-0 | .................... | .................... | ..................... | .................... | .................... | .................... |
| F | 7781 | hCoV-19/Portugal/PT1170/2020\|EPI_ISL_511716\|2020-0 | .................... | .................... | ..................... | .................... | .................... | .................... |
| F | 7782 | hCoV-19/Portugal/PT1171/2020\|EPI_ISL_511717\|2020-0 | .................... | .................... | ..................... | .................... | .................... | .................... |
| F | 7783 | hCoV-19/Portugal/PT1173/2020\|EPI_ISL_511719\|2020-0 | .................... | .................... | ..................... | .................... | .................... | .................... |
| F | 7784 | hCoV-19/Portugal/PT1174/2020\|EPI_ISL_511720\|2020-0 | .................... | .................... | ..................... | .................... | .................... | .................... |
| F | 7785 | hCoV-19/Portugal/PT1175/2020\|EPI_ISL_511721\|2020-0 | .................... | .................... | ..................... | .................... | .................... | .................... |
| F | 7786 | hCoV-19/Portugal/PT1176/2020\|EPI_ISL_511722\|2020-0 | .................... | .................... | ..................... | .................... | .................... | .................... |
| F | 7787 | hCoV-19/Portugal/PT1177/2020\|EPI_ISL_511723\|2020-0 | .................... | .................... | ..................... | .................... | .................... | .................... |
| F | 7788 | hCoV-19/Portugal/PT1178/2020\|EPI_ISL_511724\|2020-0 | .................... | .................... | ..................... | .................... | .................... | .................... |
| F | 7789 | hCoV-19/Portugal/PT1180/2020\|EPI_ISL_511725\|2020-0 | .................... | .................... | ..................... | .................... | .................... | .................... |
| F | 7790 | hCoV-19/Portugal/PT1181/2020\|EPI_ISL_511726\|2020-0 | .................... | .................... | ..................... | .................... | .................... | .................... |
| F | 7791 | hCoV-19/Portugal/PT1182/2020\|EPI_ISL_511727\|2020-0 | .................... | .................... | ..................... | .................... | .................... | .................... |
| F | 7792 | hCoV-19/Portugal/PT1183/2020\|EPI_ISL_511728\|2020-0 | .................... | .................... | ..................... | .................... | .................... | .................... |
| F | 7793 | hCoV-19/Portugal/PT1184/2020\|EPI_ISL_511729\|2020-0 | .................... | .................... | ..................... | .................... | .................... | .................... |
| F | 7794 | hCoV-19/Portugal/PT1185/2020\|EPI_ISL_511730\|2020-0 | .................... | .................... | ..................... | .................... | .................... | .................... |
| F | 7795 | hCoV-19/Canada/QC_AF7/2020\|EPI_ISL_515202\|2020-04- | .................... | .................... | ..................... | .................... | .................... | .................... |
| F | 7796 | hCoV-19/Canada/QC_AG2/2020\|EPI_ISL_515203\|2020-04- | .................... | .................... | ..................... | .................... | .................... | .................... |
| F | 7797 | hCoV-19/Canada/QC_AK6/2020\|EPI_ISL_515209\|2020-04- | .................... | .................... | ..................... | .................... | .................... | .................... |
| F | 7798 | hCoV-19/Canada/QC_AK9/2020\|EPI_ISL_515210\|2020-04- | .................... | .................... | ..................... | .................... | .................... | .................... |
| F | 7799 | hCoV-19/Canada/QC_AL3/2020\|EPI_ISL_515211\|2020-04- | .................... | .................... | ..................... | .................... | .................... | .................... |
| F | 7800 | hCoV-19/Canada/QC_AL4/2020\|EPI_ISL_515212\|2020-04- | .................... | .................... | ..................... | .................... | .................... | .................... |
| F | 7801 | hCoV-19/Canada/QC_AM4/2020\|EPI_ISL_515214\|2020-04- | .................... | .................... | ..................... | .................... | .................... | .................... |
| F | 7802 | hCoV-19/Canada/QC_AM8/2020\|EPI_ISL_515216\|2020-04- | .................... | .................... | ..................... | .................... | .................... | .................... |
| F | 7803 | hCoV-19/Canada/QC_AN6/2020\|EPI_ISL_515220\|2020-04- | .................... | .................... | ..................... | .................... | .................... | .................... |
| F | 7804 | hCoV-19/Canada/QC_AN9/2020\|EPI_ISL_515222\|2020-04- | .................... | .................... | ..................... | .................... | .................... | .................... |
| F | 7805 | hCoV-19/Canada/QC_AP4/2020\|EPI_ISL_515224\|2020-04- | .................... | .................... | ..................... | .................... | .................... | .................... |
| F | 7806 | hCoV-19/Canada/QC_AQ8/2020\|EPI_ISL_515225\|2020-04- | .................... | .................... | ..................... | .................... | .................... | .................... |
| F | 7807 | hCoV-19/Canada/QC_AR7/2020\|EPI_ISL_515226\|2020-04- | .................... | .................... | ..................... | .................... | .................... | .................... |
| F | 7808 | hCoV-19/Canada/QC_AR8/2020\|EPI_ISL_515227\|2020-04- | .................... | .................... | ..................... | .................... | .................... | .................... |
| F | 7809 | hCoV-19/Canada/QC_AT4/2020\|EPI_ISL_515228\|2020-04- | .................... | .................... | ..................... | .................... | .................... | .................... |
| F | 7810 | hCoV-19/Canada/QC_AU3/2020\|EPI_ISL_515229\|2020-04- | .................... | .................... | ..................... | .................... | .................... | .................... |
| F | 7811 | hCoV-19/Canada/QC_AU6/2020\|EPI_ISL_515230\|2020-04- | .................... | .................... | ..................... | .................... | .................... | .................... |
| F | 7812 | hCoV-19/Canada/QC_AV5/2020\|EPI_ISL_515231\|2020-04- | .................... | .................... | ..................... | .................... | .................... | .................... |
| F | 7813 | hCoV-19/Canada/QC_AW7/2020\|EPI_ISL_515233\|2020-04- | .................... | .................... | ..................... | .................... | .................... | .................... |
| F | 7814 | hCoV-19/Canada/QC_AX3/2020\|EPI_ISL_515234\|2020-04- | .................... | .................... | ..................... | .................... | .................... | .................... |
| F | 7815 | hCoV-19/Canada/QC_AY6/2020\|EPI_ISL_515235\|2020-04- | .................... | .................... | ..................... | .................... | .................... | .................... |
| F | 7816 | hCoV-19/Canada/QC_AZ6/2020\|EPI_ISL_515237\|2020-04- | .................... | .................... | ..................... | .................... | .................... | .................... |
| F | 7817 | hCoV-19/Canada/QC_BA1/2020\|EPI_ISL_515238\|2020-04- | .................... | .................... | ..................... | .................... | .................... | .................... |
| F | 7818 | hCoV-19/Canada/QC_BB3/2020\|EPI_ISL_515241\|2020-04- | .................... | .................... | ..................... | .................... | .................... | .................... |
| F | 7819 | hCoV-19/Canada/QC_BB6/2020\|EPI_ISL_515243\|2020-04- | .................... | .................... | ..................... | .................... | .................... | .................... |
| F | 7820 | hCoV-19/Canada/QC_BB9/2020\|EPI_ISL_515244\|2020-04- | .................... | .................... | ..................... | .................... | .................... | .................... |
| F | 7821 | hCoV-19/Canada/QC_BC6/2020\|EPI_ISL_515245\|2020-04- | .................... | .................... | ..................... | .................... | .................... | .................... |
| F | 7822 | hCoV-19/USA/TX-DSHS-0001/2020\|EPI_ISL_515250\|2020- | .................... | .................... | ..................... | .................... | .................... | .................... |
| F | 7823 | hCoV-19/USA/TX-DSHS-0002/2020\|EPI_ISL_515251\|2020- | .................... | .................... | ..................... | .................... | .................... | .................... |
| F | 7824 | hCoV-19/USA/TX-DSHS-0003/2020\|EPI_ISL_515252\|2020- | .................... | .................... | ..................... | .................... | .................... | .................... |
| F | 7825 | hCoV-19/USA/TX-DSHS-0004/2020\|EPI_ISL_515253\|2020- | .................... | .................... | ..................... | .................... | .................... | .................... |
| F | 7826 | hCoV-19/USA/TX-DSHS-0005/2020\|EPI_ISL_515254\|2020- | .................... | .................... | ..................... | .................... | .................... | .................... |
| F | 7827 | hCoV-19/USA/TX-DSHS-0013/2020\|EPI_ISL_515259\|2020- | .................... | .................... | ..................... | .................... | .................... | .................... |
| F | 7828 | hCoV-19/USA/MN-MDH-1454/2020\|EPI_ISL_515263\|2020-0 | .................... | .................... | ..................... | .................... | .................... | .................... |
| F | 7829 | hCoV-19/USA/MN-MDH-1456/2020\|EPI_ISL_515265\|2020-0 | .................... | .................... | ..................... | .................... | .................... | .................... |
| F | 7830 | hCoV-19/USA/MN-MDH-1457/2020\|EPI_ISL_515266\|2020-0 | .................... | .................... | ..................... | .................... | .................... | .................... |
| F | 7831 | hCoV-19/USA/ID-UW-4601/2020\|EPI_ISL_515270\|2020-03 | .................... | .................... | ..................... | .................... | .................... | .................... |
| F | 7832 | hCoV-19/USA/WA-UW-4563/2020\|EPI_ISL_515271\|2020-03 | .................... | .................... | ..................... | .................... | .................... | .................... |
| F | 7833 | hCoV-19/USA/WA-UW-4569/2020\|EPI_ISL_515272\|2020-03 | .................... | .................... | ..................... | .................... | .................... | .................... |
| F | 7834 | hCoV-19/USA/WA-UW-4581/2020\|EPI_ISL_515273\|2020-03 | .................... | .................... | ..................... | .................... | .................... | .................... |
| F | 7835 | hCoV-19/USA/WA-UW-4585/2020\|EPI_ISL_515274\|2020-03 | .................... | .................... | ..................... | .................... | .................... | .................... |
| F | 7836 | hCoV-19/USA/WA-UW-4589/2020\|EPI_ISL_515275\|2020-04 | .................... | .................... | ..................... | .................... | .................... | .................... |
| F | 7837 | hCoV-19/USA/WA-UW-4592/2020\|EPI_ISL_515276\|2020-03 | .................... | .................... | ..................... | .................... | .................... | .................... |
| F | 7838 | hCoV-19/USA/WA-UW-4599/2020\|EPI_ISL_515277\|2020-03 | .................... | .................... | ..................... | .................... | .................... | .................... |
| F | 7839 | hCoV-19/USA/WA-UW-4603/2020\|EPI_ISL_515278\|2020-03 | .................... | .................... | ..................... | .................... | .................... | .................... |
| F | 7840 | hCoV-19/USA/WA-UW-4608/2020\|EPI_ISL_515279\|2020-03 | .................... | .................... | ..................... | .................... | .................... | .................... |
| F | 7841 | hCoV-19/USA/WA-UW-4614/2020\|EPI_ISL_515280\|2020-03 | .................... | .................... | ..................... | .................... | .................... | .................... |
| F | 7842 | hCoV-19/USA/WA-UW-4615/2020\|EPI_ISL_515281\|2020-03 | .................... | .................... | ..................... | .................... | .................... | .................... |
| F | 7843 | hCoV-19/USA/WA-UW-4616/2020\|EPI_ISL_515282\|2020-03 | .................... | .................... | ..................... | .................... | .................... | .................... |
| F | 7844 | hCoV-19/USA/WA-UW-4618/2020\|EPI_ISL_515283\|2020-03 | .................... | .................... | ..................... | .................... | .................... | .................... |
| F | 7845 | hCoV-19/USA/WA-UW-4620/2020\|EPI_ISL_515284\|2020-03 | .................... | .................... | ..................... | .................... | .................... | .................... |
| F | 7846 | hCoV-19/USA/WA-UW-4623/2020\|EPI_ISL_515285\|2020-03 | .................... | .................... | ..................... | .................... | .................... | .................... |
| F | 7847 | hCoV-19/Thailand/NIH_3752/2020\|EPI_ISL_515287\|2020 | .................... | .................... | ..................... | .................... | .................... | .................... |
| F | 7848 | hCoV-19/Thailand/NIH_3623/2020\|EPI_ISL_515288\|2020 | .................... | .................... | ..................... | .................... | .................... | .................... |
| F | 7849 | hCoV-19/Thailand/NIH_3710/2020\|EPI_ISL_515289\|2020 | .................... | .................... | ..................... | .................... | .................... | .................... |

F 7850 hCoV-19/Thailand/NIH_3713/2020|EPI_ISL_515290|2020 ···················· ···················· ···················· ···················· ···················· ····················
F 7851 hCoV-19/Thailand/NIH_3744/2020|EPI_ISL_515291|2020 ···················· ···················· ···················· ···················· ···················· ····················
F 7852 hCoV-19/Thailand/NIH-3083/2020|EPI_ISL_515292|2020 ···················· ···················· ···················· ···················· ···················· ····················
F 7853 hCoV-19/USA/NV-NSPHL-A0004/2020|EPI_ISL_515293|202 ···················· ···················· ···················· ···················· ···················· ····················
F 7854 hCoV-19/USA/NV-NSPHL-A0005/2020|EPI_ISL_515294|202 ···················· ···················· ···················· ···················· ···················· ····················
F 7855 hCoV-19/USA/NV-NSPHL-A0006/2020|EPI_ISL_515295|202 ···················· ···················· ···················· ···················· ···················· ····················
F 7856 hCoV-19/USA/NV-NSPHL-A0007/2020|EPI_ISL_515296|202 ···················· ···················· ···················· ···················· ···················· ····················
F 7857 hCoV-19/USA/NV-NSPHL-A0008/2020|EPI_ISL_515297|202 ···················· ···················· ···················· ···················· ···················· ····················
F 7858 hCoV-19/USA/NV-NSPHL-A0010/2020|EPI_ISL_515299|202 ···················· ···················· ···················· ···················· ···················· ····················
F 7859 hCoV-19/USA/WI-UW-1081/2020|EPI_ISL_516507|2020-07 ···················· ···················· ···················· ···················· ···················· ····················
F 7860 hCoV-19/USA/WI-UW-1083/2020|EPI_ISL_516509|2020-07 ···················· ···················· ···················· ···················· ···················· ····················
F 7861 hCoV-19/USA/WI-UW-1084/2020|EPI_ISL_516510|2020-07 ···················· ···················· ···················· ···················· ···················· ····················
F 7862 hCoV-19/USA/IL-UW-1085/2020|EPI_ISL_516511|2020-07 ···················· ·A·················· ···················· ···················· ···················· ····················
F 7863 hCoV-19/USA/WI-UW-1086/2020|EPI_ISL_516512|2020-07 ···················· ···················· ···················· ···················· ···················· ····················
F 7864 hCoV-19/USA/WI-UW-1088/2020|EPI_ISL_516513|2020-07 ···················· ···················· ···················· ···················· ···················· ····················
F 7865 hCoV-19/USA/DC-UW-1089/2020|EPI_ISL_516514|2020-07 ···················· ·············A······ ···················· ···················· ···················· ····················
F 7866 hCoV-19/USA/WI-UW-1090/2020|EPI_ISL_516515|2020-07 ···················· ···················· ···················· ···················· ···················· ····················
F 7867 hCoV-19/USA/WI-UW-1091/2020|EPI_ISL_516516|2020-07 ···················· ···················· ···················· ···················· ···················· ····················
F 7868 hCoV-19/USA/WI-UW-1092/2020|EPI_ISL_516517|2020-07 ···················· ···················· ···················· ···················· ···················· ····················
F 7869 hCoV-19/USA/WI-UW-1093/2020|EPI_ISL_516518|2020-07 ···················· ···················· ···················· ···················· ···················· ····················
F 7870 hCoV-19/USA/WI-UW-1094/2020|EPI_ISL_516519|2020-07 ···················· ···················· ···················· ···················· ···················· ····················
F 7871 hCoV-19/USA/WI-UW-1095/2020|EPI_ISL_516520|2020-08 ···················· ···················· ···················· ···················· ···················· ····················
F 7873 hCoV-19/USA/WI-UW-1097/2020|EPI_ISL_516522|2020-07 ···················· ···················· ···················· ···················· ···················· ····················
F 7875 hCoV-19/USA/WI-UW-1099/2020|EPI_ISL_516524|2020-08 ···················· ···················· ···················· ···················· ···················· ····················
F 7876 hCoV-19/USA/WI-UW-1100/2020|EPI_ISL_516525|2020-07 ···················· ···················· ···················· ···················· ···················· ····················
F 7877 hCoV-19/USA/WI-UW-1101/2020|EPI_ISL_516526|2020-07 ···················· ·············A······ ···················· ···················· ···················· ····················
F 7878 hCoV-19/USA/WI-UW-1102/2020|EPI_ISL_516527|2020-07 ···················· ·············A······ ···················· ···················· ···················· ····················
F 7879 hCoV-19/USA/WI-UW-1103/2020|EPI_ISL_516528|2020-07 ···················· ···················· ···················· ···················· ···················· ····················
F 7880 hCoV-19/USA/WI-UW-1104/2020|EPI_ISL_516529|2020-07 ···················· ···················· ···················· ···················· ···················· ····················
F 7881 hCoV-19/USA/WI-UW-1105/2020|EPI_ISL_516530|2020-07 ···················· ··C················· ···················· ···················· ···················· ····················
F 7882 hCoV-19/USA/PN-UW-1106/2020|EPI_ISL_516531|2020-07 ···················· ···················· ···················· ···················· ···················· ····················
F 7883 hCoV-19/USA/WI-UW-1107/2020|EPI_ISL_516532|2020-07 ···················· ···················· ···················· ···················· ···················· ····················
F 7884 hCoV-19/USA/WI-UW-1108/2020|EPI_ISL_516533|2020-07 ···················· ···················· ···················· ···················· ···················· ····················
F 7885 hCoV-19/USA/WI-UW-1109/2020|EPI_ISL_516534|2020-07 ···················· ···················· ···················· ···················· ···················· ····················
F 7886 hCoV-19/USA/WI-UW-1110/2020|EPI_ISL_516535|2020-07 ···················· ···················· ···················· ···················· ···················· ····················
F 7887 hCoV-19/USA/WI-UW-1111/2020|EPI_ISL_516536|2020-07 ···················· ···················· ···················· ···················· ···················· ····················
F 7888 hCoV-19/USA/WI-UW-1112/2020|EPI_ISL_516537|2020-07 ···················· ···················· ···················· ···················· ···················· ····················
F 7889 hCoV-19/USA/WI-UW-1113/2020|EPI_ISL_516538|2020-07 ···················· ···················· ···················· ···················· ···················· ····················
F 7890 hCoV-19/USA/IL-UW-1114/2020|EPI_ISL_516539|2020-07 ···················· ···················· ···················· ···················· ···················· ····················
F 7891 hCoV-19/USA/WI-UW-1115/2020|EPI_ISL_516540|2020-07 ···················· ···················· ···················· ···················· ···················· ····················
F 7892 hCoV-19/USA/WI-UW-1116/2020|EPI_ISL_516541|2020-07 ···················· ···················· ···················· ···················· ···················· ····················
F 7893 hCoV-19/USA/WI-UW-1117/2020|EPI_ISL_516542|2020-07 ···················· ···················· ···················· ···················· ···················· ····················
F 7894 hCoV-19/USA/WI-UW-1118/2020|EPI_ISL_516543|2020-07 ···················· ···················· ···················· ···················· ···················· ····················
F 7896 hCoV-19/USA/WI-UW-1120/2020|EPI_ISL_516545|2020-07 ···················· ·A·················· ···················· ···················· ···················· ····················
F 7897 hCoV-19/USA/WI-UW-1121/2020|EPI_ISL_516546|2020-07 ···················· ···················· ···················· ···················· ···················· ····················
F 7898 hCoV-19/USA/WI-UW-1122/2020|EPI_ISL_516547|2020-07 ···················· ·A·················· ···················· ···················· ···················· ····················
F 7899 hCoV-19/USA/WI-UW-1123/2020|EPI_ISL_516548|2020-07 ···················· ···················· ···················· ···················· ···················· ····················
F 7900 hCoV-19/USA/WI-UW-1124/2020|EPI_ISL_516549|2020-07 ···················· ···················· ···················· ···················· ···················· ····················
F 7901 hCoV-19/Australia/SAP488/2020|EPI_ISL_516551|2020- ···················· ·A·················· ···················· ···················· ···················· ···A················
F 7902 hCoV-19/Switzerland/ZH-ETHZ-230000/2020|EPI_ISL_51 ···················· ···················· ···················· ···················· ···················· ····················
F 7903 hCoV-19/Switzerland/ZH-ETHZ-230002/2020|EPI_ISL_51 ···················· ···················· ···················· ···················· ···················· ····················
F 7904 hCoV-19/Switzerland/AG-ETHZ-230003/2020|EPI_ISL_51 ···················· ···················· ···················· ···················· ···················· ····················
F 7905 hCoV-19/Switzerland/AG-ETHZ-230004/2020|EPI_ISL_51 ···················· ···················· ···················· ···················· ···················· ····················
F 7906 hCoV-19/Switzerland/SZ-ETHZ-230005/2020|EPI_ISL_51 ···················· ···················· ···················· ···················· ···················· ····················
F 7907 hCoV-19/Switzerland/BL-ETHZ-230016/2020|EPI_ISL_51 ···················· ···················· ···················· ···················· ···················· ····················
F 7908 hCoV-19/Switzerland/BL-ETHZ-230017/2020|EPI_ISL_51 ···················· ·A·················· ···················· ···················· ···················· ····················
F 7909 hCoV-19/Switzerland/BL-ETHZ-230018/2020|EPI_ISL_51 ···················· ···················· ···················· ···················· ···················· ····················
F 7910 hCoV-19/Switzerland/BL-ETHZ-230019/2020|EPI_ISL_51 ···················· ···················· ···················· ···················· ···················· ····················
F 7911 hCoV-19/Switzerland/BS-ETHZ-230020/2020|EPI_ISL_51 ···················· ···················· ···················· ···················· ···················· ····················
F 7912 hCoV-19/Switzerland/BL-ETHZ-230021/2020|EPI_ISL_51 ···················· ···················· ···················· ···················· ···················· ····················
F 7913 hCoV-19/Switzerland/BL-ETHZ-230022/2020|EPI_ISL_51 ···················· ···················· ···················· ···················· ···················· ····················
F 7914 hCoV-19/Switzerland/BL-ETHZ-230023/2020|EPI_ISL_51 ···················· ···················· ···················· ···················· ···················· ····················
F 7915 hCoV-19/Switzerland/BL-ETHZ-230025/2020|EPI_ISL_51 ···················· ···················· ···················· ···················· ···················· ····················
F 7916 hCoV-19/Switzerland/BL-ETHZ-230029/2020|EPI_ISL_51 ···················· ···················· ···················· ···················· ···················· ····················
F 7917 hCoV-19/Switzerland/ZH-ETHZ-230030/2020|EPI_ISL_51 ···········T········ ···················· ···················· ···················· ···················· ····················
F 7918 hCoV-19/Switzerland/ZH-ETHZ-230031/2020|EPI_ISL_51 ···················· ···················· ···················· ···················· ···················· ····················
F 7919 hCoV-19/Switzerland/ZH-ETHZ-230032/2020|EPI_ISL_51 ···················· ···················· ···················· ···················· ···················· ····················
F 7920 hCoV-19/Switzerland/ZH-ETHZ-230033/2020|EPI_ISL_51 ···················· ···················· ···················· ···················· ···················· ····················
F 7921 hCoV-19/Switzerland/GE-ETHZ-230034/2020|EPI_ISL_51 ···················· ···················· ···················· ···················· ···················· ····················
F 7922 hCoV-19/Switzerland/ZH-ETHZ-230036/2020|EPI_ISL_51 ···················· ···················· ···················· ···················· ···················· ····················
F 7923 hCoV-19/Switzerland/ZH-ETHZ-230037/2020|EPI_ISL_51 ····S··············· ·A·················· ···················· ···················· ···················· ····················
F 7924 hCoV-19/Switzerland/BL-ETHZ-230038/2020|EPI_ISL_51 ···················· ···················· ···················· ···················· ···················· ····················
F 7925 hCoV-19/Switzerland/ZH-ETHZ-230042/2020|EPI_ISL_51 ···················· ···················· ···················· ···················· ···················· ····················
F 7926 hCoV-19/Switzerland/ZH-ETHZ-230043/2020|EPI_ISL_51 ···················· ···················· ···················· ···················· ···················· ····················
F 7927 hCoV-19/Switzerland/SZ-ETHZ-230044/2020|EPI_ISL_51 ···················· ···················· ···················· ···················· ···················· ····················
F 7928 hCoV-19/Switzerland/ZH-ETHZ-230045/2020|EPI_ISL_51 ···················· ·A·················· ···················· ···················· ···················· ····················
F 7929 hCoV-19/Switzerland/BL-ETHZ-230046/2020|EPI_ISL_51 ···················· ···················· ···················· ···················· ···················· ····················
F 7930 hCoV-19/Switzerland/BL-ETHZ-230047/2020|EPI_ISL_51 ···················· ···················· ···················· ···················· ···················· ····················
F 7931 hCoV-19/Switzerland/BL-ETHZ-230048/2020|EPI_ISL_51 ···················· ···················· ···················· ···················· ···················· ····················
F 7932 hCoV-19/Switzerland/ZH-ETHZ-230049/2020|EPI_ISL_51 ···················· ···················· ···················· ···················· ···················· ····················

| | | | | | | | | |
|---|---|---|---|---|---|---|---|---|
| F | 7933 | hCoV-19/Switzerland/BE-ETHZ-230051/2020\|EPI_ISL_51 | ···················· | ···················· | ···················· | ···················· | ···················· | ···················· |
| F | 7934 | hCoV-19/Switzerland/ZH-ETHZ-230059/2020\|EPI_ISL_51 | ···················· | ···················· | ···················· | ···················· | ···················· | ···················· |
| F | 7935 | hCoV-19/Switzerland/ZH-ETHZ-230060/2020\|EPI_ISL_51 | ···················· | ···················· | ···················· | ···················· | ···················· | ···················· |
| F | 7936 | hCoV-19/Switzerland/BE-ETHZ-230061/2020\|EPI_ISL_51 | ···················· | ···················· | ···················· | ···················· | ···················· | ···················· |
| F | 7937 | hCoV-19/South Africa/KRISP-0539/2020\|EPI_ISL_49808 | ···················· | ···················· | ···················· | ···················· | ···················· | ···················· |
| F | 7938 | hCoV-19/South Africa/KRISP-0540/2020\|EPI_ISL_49808 | ···················· | ···················· | ···················· | ···················· | ···················· | ···················· |
| F | 7939 | hCoV-19/South Africa/KRISP-0543/2020\|EPI_ISL_49808 | ···················· | ···················· | ···················· | ···················· | ···················· | ···················· |
| F | 7940 | hCoV-19/South Africa/KRISP-0544/2020\|EPI_ISL_49808 | ···················· | ···················· | ···················· | ···················· | ···················· | ···················· |
| F | 7941 | hCoV-19/South Africa/KRISP-0545/2020\|EPI_ISL_49808 | ···················· | ···················· | ···················· | ···················· | ···················· | ···················· |
| F | 7942 | hCoV-19/South Africa/KRISP-0548/2020\|EPI_ISL_49808 | ···················· | ···················· | ···················· | ···················· | ···················· | ···················· |
| F | 7943 | hCoV-19/South Africa/KRISP-0549/2020\|EPI_ISL_49808 | ···················· | ···················· | ···················· | ···················· | ···················· | ···················· |
| F | 7944 | hCoV-19/South Africa/KRISP-0550/2020\|EPI_ISL_49808 | ···················· | ···················· | ···················· | ···················· | ···················· | ···················· |
| F | 7945 | hCoV-19/Latvia/test1/2020\|EPI_ISL_498226\|2020-06-1 | ···················· | ···················· | ···················· | ···················· | ···················· | ···················· |
| F | 7946 | hCoV-19/Bangladesh/BCSIR-NILMRC-165/2020\|EPI_ISL_4 | ···················· | ···················· | ···················· | ···················· | ···················· | ···················· |
| F | 7947 | hCoV-19/Bangladesh/BCSIR-NILMRC-167/2020\|EPI_ISL_4 | ···················· | ···················· | ···················· | ···················· | ···················· | ···················· |
| F | 7948 | hCoV-19/Senegal/6626/2020\|EPI_ISL_498229\|2020-04-2 | ···················· | ···················· | ···················· | ···················· | ···················· | ···················· |
| F | 7949 | hCoV-19/Senegal/6619/2020\|EPI_ISL_498230\|2020-04-2 | ···················· | ···················· | ···················· | ···················· | ···················· | ···················· |
| F | 7950 | hCoV-19/Senegal/63263/2020\|EPI_ISL_498249\|2020-06- | ···················· | ···················· | ···················· | ···················· | ···················· | ···················· |
| F | 7951 | hCoV-19/Bangladesh/BCSIR-NILMRC-258/2020\|EPI_ISL_4 | ···················· | ···················· | ···················· | ···················· | ···················· | ···················· |
| F | 7952 | hCoV-19/Thailand/Bangkok-CONI-0192/2020\|EPI_ISL_49 | ···················· | ···················· | ···················· | ···················· | ··N················· | ···················· |
| F | 7954 | hCoV-19/Bangladesh/BCSIR-NILMRC-263/2020\|EPI_ISL_4 | ···················· | ···················· | ···················· | ···················· | ···················· | ···················· |
| F | 7955 | hCoV-19/Bangladesh/BCSIR-NILMRC-265/2020\|EPI_ISL_4 | ···················· | ···················· | ···················· | ···················· | ···················· | ···················· |
| F | 7956 | hCoV-19/Hong Kong/HKU-200723-105/2020\|EPI_ISL_4982 | ···················· | ···················· | ···················· | ···················· | ···················· | ···················· |
| F | 7957 | hCoV-19/Hong Kong/HKU-200723-106/2020\|EPI_ISL_4982 | ···················· | ···················· | ···················· | ···················· | ···················· | ···················· |
| F | 7958 | hCoV-19/Bangladesh/BCSIR-NILMRC-273/2020\|EPI_ISL_4 | ···················· | ···················· | ···················· | ···················· | ···················· | ···················· |
| F | 7959 | hCoV-19/Bangladesh/BCSIR-NILMRC-274/2020\|EPI_ISL_4 | ···················· | ···················· | ···················· | ···················· | ···················· | ···················· |
| F | 7960 | hCoV-19/USA/VA-DCLS-0680/2020\|EPI_ISL_500843\|2020- | ···················· | ···················· | ···················· | ···················· | ···················· | ···················· |
| F | 7961 | hCoV-19/USA/VA-DCLS-0682/2020\|EPI_ISL_500844\|2020- | ···················· | ···················· | ···················· | ···················· | ···················· | ···················· |
| F | 7962 | hCoV-19/USA/VA-DCLS-0684/2020\|EPI_ISL_500846\|2020- | ···················· | ···················· | ···················· | ···················· | ···················· | ···················· |
| F | 7963 | hCoV-19/USA/VA-DCLS-0685/2020\|EPI_ISL_500847\|2020- | ···················· | ···················· | ···················· | ···················· | ···················· | ···················· |
| F | 7964 | hCoV-19/USA/VA-DCLS-0686/2020\|EPI_ISL_500848\|2020- | ···················· | ···················· | ···················· | ···················· | ···················· | ···················· |
| F | 7965 | hCoV-19/USA/VA-DCLS-0687/2020\|EPI_ISL_500849\|2020- | ···················· | ···················· | ···················· | ···················· | ···················· | ···················· |
| F | 7966 | hCoV-19/USA/VA-DCLS-0689/2020\|EPI_ISL_500850\|2020- | ···················· | ···················· | ···················· | ···················· | ···················· | ···················· |
| F | 7967 | hCoV-19/USA/VA-DCLS-0690/2020\|EPI_ISL_500851\|2020- | ···················· | ···················· | ···················· | ···················· | ···················· | ···················· |
| F | 7968 | hCoV-19/USA/VA-DCLS-0691/2020\|EPI_ISL_500852\|2020- | ···················· | ···················· | ···················· | ···················· | ···················· | ···················· |
| F | 7969 | hCoV-19/USA/VA-DCLS-0692/2020\|EPI_ISL_500853\|2020- | ···················· | ···················· | ···················· | ···················· | ···················· | ···················· |
| F | 7970 | hCoV-19/Brazil/PE-IAM29/2020\|EPI_ISL_500872\|2020-0 | ···················· | ···················· | ···················· | ···················· | ···················· | ···················· |
| F | 7971 | hCoV-19/Switzerland/SG-ETHZ-190012/2020\|EPI_ISL_50 | ···················· | ···················· | ···················· | ···················· | ···················· | ···················· |
| F | 7972 | hCoV-19/Switzerland/ZH-ETHZ-190013/2020\|EPI_ISL_50 | ···················· | ···················· | ···················· | ···················· | ···················· | ···················· |
| F | 7973 | hCoV-19/Switzerland/ZH-ETHZ-190014/2020\|EPI_ISL_50 | ···················· | ···················· | ···················· | ···················· | ···················· | ···················· |
| F | 7974 | hCoV-19/Switzerland/BL-ETHZ-190016/2020\|EPI_ISL_50 | ···················· | ···················· | ···················· | ···················· | ···················· | ···················· |
| F | 7975 | hCoV-19/Switzerland/AG-ETHZ-190017/2020\|EPI_ISL_50 | ···········T········ | ···················· | ···················· | ···················· | ···················· | ···················· |
| F | 7976 | hCoV-19/Switzerland/SZ-ETHZ-190018/2020\|EPI_ISL_50 | ···················· | ···················· | ···················· | ···················· | ···················· | ···················· |
| F | 7977 | hCoV-19/Switzerland/ZH-ETHZ-190019/2020\|EPI_ISL_50 | ···················· | ·A·················· | ···················· | ···················· | ···················· | ···················· |
| F | 7978 | hCoV-19/Switzerland/GR-ETHZ-190020/2021\|EPI_ISL_50 | ···················· | ···················· | ···················· | ···················· | ···················· | ···················· |
| F | 7979 | hCoV-19/Switzerland/ZH-ETHZ-190021/2020\|EPI_ISL_50 | ···················· | ···················· | ···················· | ···················· | ···················· | ···················· |
| F | 7980 | hCoV-19/Switzerland/ZG-ETHZ-190022/2020\|EPI_ISL_50 | ···················· | ···················· | ···················· | ···················· | ···················· | ···················· |
| F | 7981 | hCoV-19/Switzerland/ZH-ETHZ-190025/2020\|EPI_ISL_50 | ···················· | ···················· | ···················· | ···················· | ···················· | ···················· |
| F | 7982 | hCoV-19/Switzerland/BE-ETHZ-190026/2020\|EPI_ISL_50 | ···················· | ···················· | ···················· | ···················· | ···················· | ···················· |
| F | 7983 | hCoV-19/Switzerland/UR-ETHZ-190027/2020\|EPI_ISL_50 | ···················· | ·A·················· | ···················· | ···················· | ···················· | ···················· |
| F | 7984 | hCoV-19/Switzerland/VS-ETHZ-190029/2020\|EPI_ISL_50 | ···················· | ···················· | ···················· | ···················· | ···················· | ···················· |
| F | 7985 | hCoV-19/Switzerland/VD-ETHZ-190030/2020\|EPI_ISL_50 | ···················· | ···················· | ···················· | ···················· | ···················· | ···················· |
| F | 7986 | hCoV-19/Switzerland/ZH-ETHZ-190031/2020\|EPI_ISL_50 | ···················· | ···················· | ···················· | ···················· | ···················· | ···················· |
| F | 7987 | hCoV-19/Switzerland/ZH-ETHZ-190032/2020\|EPI_ISL_50 | ···················· | ···················· | ···················· | ···················· | ···················· | ···················· |
| F | 7988 | hCoV-19/Switzerland/SZ-ETHZ-190033/2020\|EPI_ISL_50 | ···················· | ···················· | ···················· | ···················· | ···················· | ···················· |
| F | 7989 | hCoV-19/Switzerland/BL-ETHZ-190034/2020\|EPI_ISL_50 | ···················· | ···················· | ···················· | ···················· | ···················· | ···················· |
| F | 7990 | hCoV-19/Switzerland/ZH-ETHZ-190035/2020\|EPI_ISL_50 | ···················· | ·A·················· | ···················· | ···················· | ···················· | ···················· |
| F | 7991 | hCoV-19/Switzerland/BL-ETHZ-190036/2020\|EPI_ISL_50 | ···················· | ···················· | ···················· | ···················· | ···················· | ···················· |
| F | 7992 | hCoV-19/USA/OR-OHSU-1437/2020\|EPI_ISL_509201\|2020- | ···················· | ···················· | ···················· | ···················· | ···················· | ···················· |
| F | 7993 | hCoV-19/USA/OR-OHSU-1445/2020\|EPI_ISL_509202\|2020- | ···················· | ···················· | ···················· | ···················· | ···················· | ···················· |
| F | 7994 | hCoV-19/USA/OR-OHSU-1447/2020\|EPI_ISL_509203\|2020- | ···················· | ·A·················· | ···················· | ···················· | ···················· | ···················· |
| F | 7995 | hCoV-19/USA/OR-OHSU-1451/2020\|EPI_ISL_509204\|2020- | ···················· | ···················· | ···················· | ···················· | ···················· | ···················· |
| F | 7996 | hCoV-19/USA/OR-OHSU-1453/2020\|EPI_ISL_509205\|2020- | ···················· | ···················· | ···················· | ···················· | ···················· | ···················· |
| F | 7997 | hCoV-19/USA/OR-OHSU-1455/2020\|EPI_ISL_509206\|2020- | ···················· | ·A·················· | ···················· | ···················· | ···················· | ···················· |
| F | 7998 | hCoV-19/USA/OR-OHSU-1463/2020\|EPI_ISL_509208\|2020- | ···················· | ·A·················· | ···················· | ···················· | ···················· | ···················· |
| F | 7999 | hCoV-19/USA/OR-OHSU-1465/2020\|EPI_ISL_509209\|2020- | ···················· | ·A·················· | ···················· | ···················· | ···················· | ···················· |
| F | 8000 | hCoV-19/USA/OR-OHSU-1469/2020\|EPI_ISL_509210\|2020- | ···················· | ···················· | ···················· | ···················· | ···················· | ···················· |
| F | 8001 | hCoV-19/USA/OR-OHSU-1475/2020\|EPI_ISL_509211\|2020- | ···················· | ·A·················· | ···················· | ···················· | ···················· | ···················· |
| F | 8002 | hCoV-19/USA/OR-OHSU-1481/2020\|EPI_ISL_509212\|2020- | ···················· | ·A·················· | ···················· | ···················· | ···················· | ···················· |
| F | 8003 | hCoV-19/USA/OR-OHSU-1483/2020\|EPI_ISL_509213\|2020- | ···················· | ···················· | ···················· | ···················· | ···················· | ···················· |
| F | 8004 | hCoV-19/USA/OR-OHSU-1487/2020\|EPI_ISL_509214\|2020- | ···················· | ·A·················· | ···················· | ···················· | ···················· | ···················· |
| F | 8005 | hCoV-19/Switzerland/ZH-UZH-1000503309/2020\|EPI_ISL | ···················· | ···················· | ···················· | ···················· | ···················· | ···················· |
| F | 8006 | hCoV-19/South Africa/KRISP-K000566/2020\|EPI_ISL_50 | ···················· | ···················· | ···················· | ···················· | ···················· | ···················· |
| F | 8007 | hCoV-19/South Africa/KRISP-K000567/2020\|EPI_ISL_50 | ···················· | ···················· | ···················· | ···················· | ···················· | ···················· |
| F | 8008 | hCoV-19/South Africa/KRISP-K000568/2020\|EPI_ISL_50 | ···················· | ···················· | ···················· | ···················· | ···················· | ···················· |
| F | 8009 | hCoV-19/South Africa/KRISP-K000569/2020\|EPI_ISL_50 | ···················· | ···················· | ···················· | ···················· | ···················· | ···················· |
| F | 8010 | hCoV-19/South Africa/KRISP-K000572/2020\|EPI_ISL_50 | ···················· | ···················· | ···················· | ···················· | ···················· | ···················· |
| F | 8011 | hCoV-19/South Africa/KRISP-K000573/2020\|EPI_ISL_50 | ···················· | ···················· | ···················· | ···················· | ···················· | ···················· |
| F | 8012 | hCoV-19/South Africa/KRISP-K000574/2020\|EPI_ISL_50 | ···················· | ···················· | ···················· | ···················· | ···················· | ···················· |
| F | 8013 | hCoV-19/South Africa/KRISP-K000575/2020\|EPI_ISL_50 | ···················· | ···················· | ···················· | ···················· | ···················· | ···················· |

| | | | | | | | | |
|---|---|---|---|---|---|---|---|---|
| F | 8014 | hCoV-19/South Africa/KRISP-K000576/2020\|EPI_ISL_50 | ···················· | ···················· | ····················· | ···················· | ···················· | ···················· |
| F | 8015 | hCoV-19/South Africa/KRISP-K000577/2020\|EPI_ISL_50 | ···················· | ···················· | ····················· | ···················· | ···················· | ···················· |
| F | 8016 | hCoV-19/South Africa/KRISP-K000578/2020\|EPI_ISL_50 | ···················· | ···················· | ····················· | ···················· | ···················· | ···················· |
| F | 8017 | hCoV-19/South Africa/KRISP-K000579/2020\|EPI_ISL_50 | ···················· | ···················· | ····················· | ···················· | ···················· | ···················· |
| F | 8018 | hCoV-19/South Africa/KRISP-K000580/2020\|EPI_ISL_50 | ···················· | ···················· | ····················· | ···················· | ···················· | ···················· |
| F | 8019 | hCoV-19/South Africa/KRISP-K000581/2020\|EPI_ISL_50 | ···················· | ···················· | ····················· | ···················· | ···················· | ···················· |
| F | 8020 | hCoV-19/South Africa/KRISP-K000582/2020\|EPI_ISL_50 | ···················· | ···················· | ····················· | ···················· | ···················· | ···················· |
| F | 8021 | hCoV-19/South Africa/KRISP-K000583/2020\|EPI_ISL_50 | ···················· | ···················· | ····················· | ···················· | ···················· | ···················· |
| F | 8022 | hCoV-19/South Africa/KRISP-K000584/2020\|EPI_ISL_50 | ···················· | ···················· | ····················· | ···················· | ···················· | ···················· |
| F | 8023 | hCoV-19/South Africa/KRISP-K000585/2020\|EPI_ISL_50 | ···················· | ···················· | ····················· | ···················· | ···················· | ···················· |
| F | 8025 | hCoV-19/South Africa/KRISP-K000588/2020\|EPI_ISL_50 | ···················· | ···················· | ····················· | ···················· | ···················· | ···················· |
| F | 8026 | hCoV-19/South Africa/KRISP-K000589/2020\|EPI_ISL_50 | ···················· | ···················· | ····················· | ···················· | ···················· | ···················· |
| F | 8027 | hCoV-19/South Africa/KRISP-K000591/2020\|EPI_ISL_50 | ···················· | ···················· | ····················· | ···················· | ···················· | ···················· |
| F | 8028 | hCoV-19/South Africa/KRISP-K000592/2020\|EPI_ISL_50 | ···················· | ···················· | ····················· | ···················· | ···················· | ···················· |
| F | 8029 | hCoV-19/South Africa/KRISP-K000594/2020\|EPI_ISL_50 | ···················· | ···················· | ····················· | ···················· | ···················· | ···················· |
| F | 8030 | hCoV-19/South Africa/KRISP-K000595/2020\|EPI_ISL_50 | ···················· | ···················· | ····················· | ···················· | ···················· | ···················· |
| F | 8031 | hCoV-19/South Africa/KRISP-K000596/2020\|EPI_ISL_50 | ···················· | ···················· | ····················· | ···················· | ···················· | ···················· |
| F | 8032 | hCoV-19/South Africa/KRISP-K000597/2020\|EPI_ISL_50 | ···················· | ···················· | ····················· | ···················· | ···················· | ···················· |
| F | 8033 | hCoV-19/South Africa/KRISP-K000598/2020\|EPI_ISL_50 | ···················· | ···················· | ····················· | ···················· | ···················· | ···················· |
| F | 8034 | hCoV-19/South Africa/KRISP-K000599/2020\|EPI_ISL_50 | ···················· | ···················· | ····················· | ···················· | ···················· | ···················· |
| F | 8035 | hCoV-19/South Africa/KRISP-K000600/2020\|EPI_ISL_50 | ···················· | ···················· | ····················· | ···················· | ···················· | ···················· |
| F | 8036 | hCoV-19/South Africa/KRISP-K000601/2020\|EPI_ISL_50 | ···················· | ···················· | ····················· | ···················· | ··T················· | ···················· |
| F | 8037 | hCoV-19/South Africa/KRISP-K000602/2020\|EPI_ISL_50 | ···················· | ···················· | ····················· | ···················· | ···················· | ···················· |
| F | 8038 | hCoV-19/South Africa/KRISP-K000603/2020\|EPI_ISL_50 | ···················· | ···················· | ····················· | ···················· | ···················· | ···················· |
| F | 8039 | hCoV-19/South Africa/KRISP-K000604/2020\|EPI_ISL_50 | ···················· | ···················· | ····················· | ···················· | ···················· | ···················· |
| F | 8040 | hCoV-19/South Africa/KRISP-K000605/2020\|EPI_ISL_50 | ···················· | ···················· | ····················· | ···················· | ···················· | ···················· |
| F | 8055 | hCoV-19/Spain/MD-IBV-005345/2020\|EPI_ISL_510222\|20 | ···················· | ···················· | ····················· | ···················· | ···················· | ···················· |
| F | 8072 | hCoV-19/Portugal/PT0952/2020\|EPI_ISL_511502\|2020-0 | ···················· | ······A············· | ····················· | ···················· | ···················· | ···················· |
| F | 8073 | hCoV-19/Portugal/PT0953/2020\|EPI_ISL_511503\|2020-0 | ···················· | ···················· | ····················· | ···················· | ···················· | ···················· |
| F | 8074 | hCoV-19/Portugal/PT0955/2020\|EPI_ISL_511504\|2020-0 | ···················· | ···················· | ····················· | ···················· | ···················· | ···················· |
| F | 8075 | hCoV-19/Portugal/PT0956/2020\|EPI_ISL_511505\|2020-0 | ···················· | ···················· | ····················· | ···················· | ···················· | ···················· |
| F | 8076 | hCoV-19/Portugal/PT0957/2020\|EPI_ISL_511506\|2020-0 | ···················· | ···················· | ····················· | ···················· | ···················· | ···················· |
| F | 8077 | hCoV-19/Portugal/PT0958/2020\|EPI_ISL_511507\|2020-0 | ···················· | ···················· | ····················· | ···················· | ···················· | ···················· |
| F | 8078 | hCoV-19/Portugal/PT0959/2020\|EPI_ISL_511508\|2020-0 | ···················· | ···················· | ····················· | ···················· | ···················· | ···················· |
| F | 8079 | hCoV-19/Portugal/PT0960/2020\|EPI_ISL_511509\|2020-0 | ···················· | ···················· | ····················· | ···················· | ···················· | ···················· |
| F | 8080 | hCoV-19/Portugal/PT0961/2020\|EPI_ISL_511510\|2020-0 | ···················· | ···················· | ····················· | ···················· | ···················· | ···················· |
| F | 8081 | hCoV-19/Portugal/PT0963/2020\|EPI_ISL_511512\|2020-0 | ···················· | ···················· | ····················· | ···················· | ···················· | ···················· |
| F | 8082 | hCoV-19/Portugal/PT0964/2020\|EPI_ISL_511513\|2020-0 | ···················· | ···················· | ····················· | ···················· | ···················· | ···················· |
| F | 8083 | hCoV-19/Portugal/PT0965/2020\|EPI_ISL_511514\|2020-0 | ···················· | ···················· | ····················· | ···················· | ···················· | ···················· |
| F | 8084 | hCoV-19/Portugal/PT0966/2020\|EPI_ISL_511515\|2020-0 | ···················· | ···················· | ····················· | ···················· | ···················· | ···················· |
| F | 8085 | hCoV-19/Portugal/PT0967/2020\|EPI_ISL_511516\|2020-0 | ···················· | ···················· | ····················· | ···················· | ···················· | ···················· |
| F | 8086 | hCoV-19/Portugal/PT0968/2020\|EPI_ISL_511517\|2020-0 | ···················· | ···················· | ····················· | ···················· | ···················· | ···················· |
| F | 8087 | hCoV-19/Portugal/PT0969/2020\|EPI_ISL_511518\|2020-0 | ···················· | ···················· | ····················· | ···················· | ···················· | ···················· |
| F | 8088 | hCoV-19/Portugal/PT0971/2020\|EPI_ISL_511520\|2020-0 | ···················· | ···················· | ····················· | ···················· | ···················· | ···················· |
| F | 8089 | hCoV-19/Portugal/PT0972/2020\|EPI_ISL_511521\|2020-0 | ···················· | ···················· | ····················· | ···················· | ···················· | ···················· |
| F | 8090 | hCoV-19/Portugal/PT0973/2020\|EPI_ISL_511522\|2020-0 | ···················· | ···················· | ····················· | ···················· | ···················· | ···················· |
| F | 8091 | hCoV-19/Portugal/PT0974/2020\|EPI_ISL_511523\|2020-0 | ···················· | ···················· | ····················· | ···················· | ···················· | ···················· |
| F | 8092 | hCoV-19/Portugal/PT0975/2020\|EPI_ISL_511524\|2020-0 | ···················· | ···················· | ····················· | ···················· | ···················· | ···················· |
| F | 8093 | hCoV-19/Portugal/PT0976/2020\|EPI_ISL_511525\|2020-0 | ···················· | ···················· | ····················· | ···················· | ···················· | ···················· |
| F | 8094 | hCoV-19/Portugal/PT0977/2020\|EPI_ISL_511526\|2020-0 | ···················· | ···················· | ····················· | ···················· | ···················· | ···················· |
| F | 8095 | hCoV-19/Portugal/PT0978/2020\|EPI_ISL_511527\|2020-0 | ···················· | ···················· | ····················· | ···················· | ···················· | ···················· |
| F | 8096 | hCoV-19/Portugal/PT0979/2020\|EPI_ISL_511528\|2020-0 | ···················· | ···················· | ····················· | ···················· | ···················· | ···················· |
| F | 8097 | hCoV-19/Portugal/PT0980/2020\|EPI_ISL_511529\|2020-0 | ···················· | ···················· | ····················· | ···················· | ···················· | ···················· |
| F | 8098 | hCoV-19/Portugal/PT0981/2020\|EPI_ISL_511530\|2020-0 | ···················· | ···················· | ····················· | ···················· | ···················· | ···················· |
| F | 8099 | hCoV-19/Portugal/PT0983/2020\|EPI_ISL_511531\|2020-0 | ···················· | ···················· | ····················· | ···················· | ···················· | ···················· |
| F | 8100 | hCoV-19/Portugal/PT0984/2020\|EPI_ISL_511532\|2020-0 | ···················· | ···················· | ····················· | ···················· | ···················· | ···················· |
| F | 8101 | hCoV-19/Portugal/PT0985/2020\|EPI_ISL_511533\|2020-0 | ···················· | ···················· | ····················· | ···················· | ···················· | ···················· |
| F | 8102 | hCoV-19/Portugal/PT0986/2020\|EPI_ISL_511534\|2020-0 | ···················· | ···················· | ····················· | ···················· | ···················· | ···················· |
| F | 8103 | hCoV-19/Portugal/PT0987/2020\|EPI_ISL_511535\|2020-0 | ···················· | ···················· | ····················· | ···················· | ···················· | ···················· |
| F | 8104 | hCoV-19/Portugal/PT0988/2020\|EPI_ISL_511536\|2020-0 | ···················· | ···················· | ····················· | ···················· | ···················· | ···················· |
| F | 8105 | hCoV-19/Portugal/PT0989/2020\|EPI_ISL_511537\|2020-0 | ···················· | ···················· | ····················· | ···················· | ···················· | ···················· |
| F | 8106 | hCoV-19/Portugal/PT0991/2020\|EPI_ISL_511539\|2020-0 | ···················· | ···················· | ····················· | ···················· | ···················· | ···················· |
| F | 8107 | hCoV-19/Portugal/PT0993/2020\|EPI_ISL_511541\|2020-0 | ···················· | ···················· | ····················· | ···················· | ···················· | ···················· |
| F | 8108 | hCoV-19/Portugal/PT0994/2020\|EPI_ISL_511542\|2020-0 | ···················· | ···················· | ····················· | ···················· | ···················· | ···················· |
| F | 8109 | hCoV-19/Portugal/PT0995/2020\|EPI_ISL_511543\|2020-0 | ···················· | ···················· | ····················· | ···················· | ···················· | ···················· |
| F | 8110 | hCoV-19/Portugal/PT0996/2020\|EPI_ISL_511544\|2020-0 | ···················· | ···················· | ····················· | ···················· | ···················· | ···················· |
| F | 8111 | hCoV-19/Portugal/PT0997/2020\|EPI_ISL_511545\|2020-0 | ···················· | ···················· | ····················· | ···················· | ···················· | ···················· |
| F | 8112 | hCoV-19/Portugal/PT0998/2020\|EPI_ISL_511546\|2020-0 | ···················· | ···················· | ····················· | ···················· | ···················· | ···················· |
| F | 8113 | hCoV-19/Portugal/PT1001/2020\|EPI_ISL_511549\|2020-0 | ···················· | ···················· | ····················· | ···················· | ···················· | ···················· |
| F | 8114 | hCoV-19/Portugal/PT1004/2020\|EPI_ISL_511551\|2020-0 | ···················· | ···················· | ····················· | ···················· | ···················· | ···················· |
| F | 8115 | hCoV-19/Portugal/PT1006/2020\|EPI_ISL_511553\|2020-0 | ···················· | ···················· | ····················· | ···················· | ···················· | ···················· |
| F | 8116 | hCoV-19/Portugal/PT1009/2020\|EPI_ISL_511556\|2020-0 | ···················· | ···················· | ····················· | ···················· | ···················· | ···················· |
| F | 8117 | hCoV-19/Portugal/PT1010/2020\|EPI_ISL_511557\|2020-0 | ···················· | ···················· | ····················· | ···················· | ···················· | ···················· |
| F | 8118 | hCoV-19/Portugal/PT1011/2020\|EPI_ISL_511558\|2020-0 | ···················· | ···················· | ····················· | ···················· | ···················· | ···················· |
| F | 8119 | hCoV-19/Portugal/PT1012/2020\|EPI_ISL_511559\|2020-0 | ···················· | ···················· | ····················· | ···················· | ···················· | ···················· |
| F | 8120 | hCoV-19/Portugal/PT1013/2020\|EPI_ISL_511560\|2020-0 | ···················· | ···················· | ····················· | ···················· | ···················· | ···················· |
| F | 8121 | hCoV-19/Portugal/PT1014/2020\|EPI_ISL_511561\|2020-0 | ···················· | ···················· | ····················· | ···················· | ···················· | ···················· |
| F | 8122 | hCoV-19/Portugal/PT1016/2020\|EPI_ISL_511563\|2020-0 | ···················· | ···················· | ····················· | ···················· | ···················· | ···················· |
| F | 8123 | hCoV-19/Portugal/PT1018/2020\|EPI_ISL_511565\|2020-0 | ···················· | ···················· | ····················· | ···················· | ···················· | ···················· |
| F | 8124 | hCoV-19/Portugal/PT1019/2020\|EPI_ISL_511566\|2020-0 | ···················· | ···················· | ····················· | ···················· | ···················· | ···················· |

| | | | | | | | |
|---|---|---|---|---|---|---|---|
| F | 8125 hCoV-19/Portugal/PT1020/2020\|EPI_ISL_511567\|2020-0 | ···················· | ···················· | ···················· | ···················· | ···················· | ···················· |
| F | 8126 hCoV-19/Portugal/PT1022/2020\|EPI_ISL_511569\|2020-0 | ···················· | ···················· | ···················· | ···················· | ···················· | ···················· |
| F | 8127 hCoV-19/Portugal/PT1023/2020\|EPI_ISL_511570\|2020-0 | ···················· | ···················· | ···················· | ···················· | ···················· | ···················· |
| F | 8128 hCoV-19/Portugal/PT1024/2020\|EPI_ISL_511571\|2020-0 | ···················· | ···················· | ···················· | ···················· | ···················· | ···················· |
| F | 8129 hCoV-19/Portugal/PT1029/2020\|EPI_ISL_511576\|2020-0 | ···················· | ···················· | ···················· | ···················· | ···················· | ···················· |
| F | 8130 hCoV-19/Singapore/797/2020\|EPI_ISL_512833\|2020-07- | ···················· | ···················· | ···················· | ···················· | ···················· | ···················· |
| F | 8131 hCoV-19/Singapore/798/2020\|EPI_ISL_512834\|2020-07- | ···················· | ···················· | ···················· | ···················· | ···················· | ···················· |
| F | 8132 hCoV-19/Singapore/799/2020\|EPI_ISL_512835\|2020-07- | ···················· | ·A·················· | ···················· | ···················· | ···················· | ··A················· |
| F | 8133 hCoV-19/Singapore/800/2020\|EPI_ISL_512836\|2020-07- | ···················· | ·A·················· | ···················· | ···················· | ···················· | ··A················· |
| F | 8134 hCoV-19/Singapore/801/2020\|EPI_ISL_512837\|2020-07- | ···················· | ···················· | ···················· | ···················· | ···················· | ···················· |
| F | 8135 hCoV-19/Singapore/802/2020\|EPI_ISL_512838\|2020-07- | ···················· | ···················· | ···················· | ···················· | ···················· | ···················· |
| F | 8136 hCoV-19/Singapore/803/2020\|EPI_ISL_512839\|2020-07- | ···················· | ···················· | ···················· | ···················· | ···················· | ···················· |
| F | 8137 hCoV-19/Singapore/804/2020\|EPI_ISL_512840\|2020-07- | ···················· | ···················· | ···················· | ···················· | ···················· | ···················· |
| F | 8138 hCoV-19/Singapore/805/2020\|EPI_ISL_512841\|2020-07- | ···················· | ···················· | ···················· | ···················· | ···················· | ···················· |
| F | 8139 hCoV-19/Singapore/806/2020\|EPI_ISL_512842\|2020-08- | ···················· | ···················· | ···················· | ···················· | ···················· | ···················· |
| F | 8140 hCoV-19/Myanmar/MMC_137/2020\|EPI_ISL_512844\|2020-0 | ···················· | ···················· | ···················· | ···················· | ···················· | ···················· |
| F | 8141 hCoV-19/Costa Rica/INC-0066/2020\|EPI_ISL_512846\|20 | ···················· | ···················· | ···················· | ···················· | ···················· | ···················· |
| F | 8144 hCoV-19/Thailand/Bangkok-CONI-0202/2020\|EPI_ISL_51 | ···················· | ···················· | ···················· | ···················· | ···················· | ···················· |
| F | 8146 hCoV-19/Thailand/Bangkok-CONI-0207/2020\|EPI_ISL_51 | ···················· | ···················· | ···················· | ···················· | ···················· | ···················· |
| F | 8149 hCoV-19/Thailand/Bangkok-CONI-0210/2020\|EPI_ISL_51 | ···················· | ···················· | ···················· | ···················· | ···················· | ···················· |
| F | 8150 hCoV-19/Thailand/Bangkok-CONI-0211/2020\|EPI_ISL_51 | ···················· | ···················· | ···················· | ···················· | ···················· | ···················· |
| F | 8151 hCoV-19/Thailand/Bangkok-CONI-0212/2020\|EPI_ISL_51 | ···················· | ···················· | ···················· | ···················· | ···················· | ···················· |
| F | 8152 hCoV-19/Thailand/Bangkok-CONI-0213/2020\|EPI_ISL_51 | ···················· | ···················· | ···················· | ···················· | ···················· | ···················· |
| F | 8153 hCoV-19/Thailand/Bangkok-CONI-0214/2020\|EPI_ISL_51 | ···················· | ···················· | ···················· | ···················· | ···················· | ···················· |
| F | 8154 hCoV-19/Thailand/Bangkok-CONI-0224/2020\|EPI_ISL_51 | ···················· | ···················· | ···················· | ···················· | ···················· | ···················· |
| F | 8155 hCoV-19/Thailand/Bangkok-CONI-0227/2020\|EPI_ISL_51 | ···················· | ···················· | ···················· | ···················· | ···················· | ···················· |
| F | 8156 hCoV-19/SaudiArabia/KAUST-JEDDAH1000/2020\|EPI_ISL_ | ···················· | ···················· | ···················· | ···················· | ···················· | ···················· |
| F | 8157 hCoV-19/SaudiArabia/KAUST-JEDDAH1001/2020\|EPI_ISL_ | ···················· | ···················· | ···················· | ···················· | ···················· | ···················· |
| F | 8158 hCoV-19/SaudiArabia/KAUST-JEDDAH1002/2020\|EPI_ISL_ | ···················· | ···················· | ···················· | ···················· | ···················· | ···················· |
| F | 8159 hCoV-19/SaudiArabia/KAUST-JEDDAH1003/2020\|EPI_ISL_ | ···················· | ···················· | ···················· | ···················· | ···················· | ···················· |
| F | 8160 hCoV-19/SaudiArabia/KAUST-JEDDAH1007/2020\|EPI_ISL_ | ···················· | ·A·················· | ···················· | ···················· | ···················· | ··A················· |
| F | 8161 hCoV-19/SaudiArabia/KAUST-JEDDAH1008/2020\|EPI_ISL_ | ···················· | ·A·················· | ···················· | ···················· | ···················· | ··A················· |
| F | 8162 hCoV-19/SaudiArabia/KAUST-JEDDAH1009/2020\|EPI_ISL_ | ···················· | ···················· | ···················· | ···················· | ···················· | ···················· |
| F | 8163 hCoV-19/SaudiArabia/KAUST-JEDDAH1010/2020\|EPI_ISL_ | ···················· | ···················· | ···················· | ···················· | ···················· | ···················· |
| F | 8164 hCoV-19/SaudiArabia/KAUST-JEDDAH1011/2020\|EPI_ISL_ | ···················· | ·A·················· | ···················· | ···················· | ···················· | ··A················· |
| F | 8165 hCoV-19/SaudiArabia/KAUST-JEDDAH1012/2020\|EPI_ISL_ | ···················· | ···················· | ···················· | ···················· | ···················· | ···················· |
| F | 8166 hCoV-19/SaudiArabia/KAUST-JEDDAH1013/2020\|EPI_ISL_ | ···················· | ···················· | ···················· | ···················· | ···················· | ···················· |
| F | 8167 hCoV-19/SaudiArabia/KAUST-JEDDAH1014/2020\|EPI_ISL_ | ···················· | ···················· | ···················· | ···················· | ···················· | ···················· |
| F | 8168 hCoV-19/SaudiArabia/KAUST-JEDDAH1015/2020\|EPI_ISL_ | ···················· | ···················· | ···················· | ···················· | ···················· | ···················· |
| F | 8169 hCoV-19/SaudiArabia/KAUST-JEDDAH1016/2020\|EPI_ISL_ | ···················· | ···················· | ···················· | ···················· | ···················· | ···················· |
| F | 8170 hCoV-19/SaudiArabia/KAUST-JEDDAH1017/2020\|EPI_ISL_ | ···················· | ·A·················· | ···················· | ···················· | ···················· | ··A················· |
| F | 8171 hCoV-19/SaudiArabia/KAUST-JEDDAH1018/2020\|EPI_ISL_ | ···················· | ···················· | ···················· | ···················· | ···················· | ···················· |
| F | 8172 hCoV-19/SaudiArabia/KAUST-JEDDAH1020/2020\|EPI_ISL_ | ···················· | ···················· | ···················· | ···················· | ···················· | ···················· |
| F | 8173 hCoV-19/SaudiArabia/KAUST-JEDDAH1021/2020\|EPI_ISL_ | ···················· | ···················· | ···················· | ···················· | ···················· | ···················· |
| F | 8174 hCoV-19/SaudiArabia/KAUST-JEDDAH1022/2020\|EPI_ISL_ | ···················· | ···················· | ···················· | ···················· | ···················· | ···················· |
| F | 8175 hCoV-19/SaudiArabia/KAUST-JEDDAH1023/2020\|EPI_ISL_ | ···················· | ···················· | ···················· | ···················· | ···················· | ···················· |
| F | 8176 hCoV-19/SaudiArabia/KAUST-JEDDAH1024/2020\|EPI_ISL_ | ···················· | ···················· | ···················· | ···················· | ···················· | ···················· |
| F | 8177 hCoV-19/SaudiArabia/KAUST-JEDDAH1025/2020\|EPI_ISL_ | ···················· | ···················· | ···················· | ···················· | ···················· | ···················· |
| F | 8178 hCoV-19/SaudiArabia/KAUST-JEDDAH1026/2020\|EPI_ISL_ | ···················· | ···················· | ···················· | ···················· | ···················· | ···················· |
| F | 8179 hCoV-19/SaudiArabia/KAUST-JEDDAH1028/2020\|EPI_ISL_ | ···················· | ···················· | ···················· | ···················· | ···················· | ···················· |
| F | 8180 hCoV-19/SaudiArabia/KAUST-JEDDAH1030/2020\|EPI_ISL_ | ···················· | ···················· | ···················· | ···················· | ···················· | ···················· |
| F | 8181 hCoV-19/USA/MN-MDH-1364/2020\|EPI_ISL_515262\|2020-0 | ···················· | ···················· | ···················· | ···················· | ···················· | ···················· |
| F | 8182 hCoV-19/USA/MN-MDH-1455/2020\|EPI_ISL_515264\|2020-0 | ···················· | ···················· | ···················· | ···················· | ···················· | ···················· |
| F | 8183 hCoV-19/USA/MN-MDH-1458/2020\|EPI_ISL_515267\|2020-0 | ···················· | ···················· | ···················· | ···················· | ···················· | ···················· |
| F | 8184 hCoV-19/USA/MN-MDH-1459/2020\|EPI_ISL_515268\|2020-0 | ···················· | ···················· | ···················· | ···················· | ···················· | ···················· |
| F | 8185 hCoV-19/USA/MN-MDH-1460/2020\|EPI_ISL_515269\|2020-0 | ···················· | ···················· | ···················· | ···················· | ···················· | ···················· |
| F | 8186 hCoV-19/Switzerland/BE-ETHZ-230052/2020\|EPI_ISL_51 | ···················· | ···················· | ···················· | ···················· | ···················· | ···················· |
| F | 8187 hCoV-19/Switzerland/BE-ETHZ-230053/2020\|EPI_ISL_51 | ···················· | ···················· | ···················· | ···················· | ···················· | ···················· |
| F | 8188 hCoV-19/Switzerland/ZH-ETHZ-230055/2020\|EPI_ISL_51 | ···················· | ···················· | ···················· | ···················· | ···················· | ···················· |
| F | 8189 hCoV-19/Switzerland/ZH-ETHZ-230057/2020\|EPI_ISL_51 | ···················· | ···················· | ···················· | ···················· | ···················· | ···················· |
| F | 8190 hCoV-19/Switzerland/ZH-ETHZ-230058/2020\|EPI_ISL_51 | ···················· | ···················· | ···················· | ···················· | ···················· | ···················· |
| F | 8191 hCoV-19/Switzerland/BE-ETHZ-230062/2020\|EPI_ISL_51 | ···················· | ·A·················· | ···················· | ···················· | ···················· | ···················· |
| F | 8192 hCoV-19/Switzerland/VD-ETHZ-230064/2020\|EPI_ISL_51 | ···················· | ···················· | ···················· | ···················· | ···················· | ···················· |
| F | 8193 hCoV-19/USA/WA-S1514/2020\|EPI_ISL_495940\|2020-05-2 | ···················· | ···················· | ···················· | ···················· | ···················· | ···················· |
| F | 8194 hCoV-19/USA/WA-S1517/2020\|EPI_ISL_495943\|2020-05-2 | ···················· | ···················· | ···················· | ···················· | ···················· | ···················· |
| F | 8195 hCoV-19/USA/WA-S1518/2020\|EPI_ISL_495944\|2020-05-2 | ···················· | ···················· | ···················· | ···················· | ···················· | ···················· |
| F | 8196 hCoV-19/USA/WA-S1520/2020\|EPI_ISL_495946\|2020-05-2 | ···················· | ···················· | ···················· | ···················· | ···················· | ···················· |
| F | 8197 hCoV-19/USA/WA-S1521/2020\|EPI_ISL_495947\|2020-05-2 | ···················· | ···················· | ···················· | ···················· | ···················· | ···················· |
| F | 8198 hCoV-19/USA/WA-S1522/2020\|EPI_ISL_495948\|2020-05-2 | ···················· | ···················· | ···················· | ···················· | ···················· | ···················· |
| F | 8199 hCoV-19/USA/WA-S1523/2020\|EPI_ISL_495949\|2020-05-2 | ···················· | ··············N····· | ···················· | ···················· | ···················· | ···················· |
| F | 8200 hCoV-19/USA/WA-S1524/2020\|EPI_ISL_495950\|2020-05-2 | ···················· | ···················· | ···················· | ···················· | ···················· | ···················· |
| F | 8201 hCoV-19/USA/WA-S1525/2020\|EPI_ISL_495951\|2020-05-2 | ···················· | ···················· | ···················· | ···················· | ···················· | ···················· |
| F | 8202 hCoV-19/USA/WA-S1526/2020\|EPI_ISL_495952\|2020-05-2 | ···················· | ···················· | ···················· | ···················· | ···················· | ···················· |
| F | 8203 hCoV-19/USA/WA-S1528/2020\|EPI_ISL_495954\|2020-05-2 | ···················· | ···················· | ···················· | ···················· | ···················· | ···················· |
| F | 8204 hCoV-19/USA/WA-S1529/2020\|EPI_ISL_495955\|2020-05-2 | ···················· | ···················· | ···················· | ···················· | ···················· | ···················· |
| F | 8205 hCoV-19/USA/WA-S1530/2020\|EPI_ISL_495956\|2020-05-2 | ···················· | ···················· | ···················· | ···················· | ···················· | ···················· |
| F | 8206 hCoV-19/USA/WA-S1533/2020\|EPI_ISL_495959\|2020-05-2 | ···················· | ···················· | ···················· | ···················· | ···················· | ···················· |
| F | 8207 hCoV-19/USA/WA-S1544/2020\|EPI_ISL_495970\|2020-05-2 | ···················· | ···················· | ···················· | ···················· | ···················· | ···················· |
| F | 8208 hCoV-19/USA/WA-S1546/2020\|EPI_ISL_495972\|2020-05-2 | ···················· | ···················· | ···················· | ···················· | ···················· | ···················· |
| F | 8209 hCoV-19/USA/WA-S1547/2020\|EPI_ISL_495973\|2020-05-2 | ···················· | ···················· | ···················· | ···················· | ···················· | ···················· |

| | | | | | | | | |
|---|---|---|---|---|---|---|---|---|
| F | 8210 | hCoV-19/USA/WA-S1548/2020\|EPI_ISL_495974\|2020-05-2 | ···················· | ···················· | ····················· | ···················· | ···················· | ···················· |
| F | 8211 | hCoV-19/USA/WA-S1549/2020\|EPI_ISL_495975\|2020-05-2 | ···················· | ···················· | ····················· | ···················· | ···················· | ···················· |
| F | 8212 | hCoV-19/USA/WA-S1551/2020\|EPI_ISL_495977\|2020-05-2 | ···················· | ···················· | ····················· | ···················· | ···················· | ···················· |
| F | 8213 | hCoV-19/USA/WA-S1555/2020\|EPI_ISL_495981\|2020-05-2 | ···················· | ···················· | ····················· | ···················· | ···················· | ···················· |
| F | 8214 | hCoV-19/USA/WA-S1556/2020\|EPI_ISL_495982\|2020-05-2 | ···················· | ···················· | ····················· | ···················· | ···················· | ···················· |
| F | 8215 | hCoV-19/USA/WA-S1557/2020\|EPI_ISL_495983\|2020-05-2 | ···················· | ···················· | ····················· | ···················· | ···················· | ···················· |
| F | 8216 | hCoV-19/USA/WA-S1558/2020\|EPI_ISL_495984\|2020-05-2 | ···················· | ···················· | ····················· | ···················· | ···················· | ···················· |
| F | 8217 | hCoV-19/USA/WA-S1559/2020\|EPI_ISL_495985\|2020-05-2 | ···················· | ···················· | ····················· | ···················· | ···················· | ···················· |
| F | 8218 | hCoV-19/USA/WA-S1560/2020\|EPI_ISL_495986\|2020-05-2 | ···················· | ···················· | ····················· | ···················· | ···················· | ···················· |
| F | 8219 | hCoV-19/USA/WA-S1561/2020\|EPI_ISL_495987\|2020-05-2 | ···················· | ···················· | ····················· | ···················· | ···················· | ···················· |
| F | 8220 | hCoV-19/USA/WA-S1562/2020\|EPI_ISL_495988\|2020-05-2 | ···················· | ···················· | ····················· | ···················· | ···················· | ···················· |
| F | 8221 | hCoV-19/USA/WA-S1563/2020\|EPI_ISL_495989\|2020-05-2 | ···················· | ···················· | ····················· | ···················· | ···················· | ···················· |
| F | 8222 | hCoV-19/USA/WA-S1565/2020\|EPI_ISL_495991\|2020-05-2 | ···················· | ···················· | ····················· | ···················· | ···················· | ···················· |
| F | 8223 | hCoV-19/USA/WA-S1566/2020\|EPI_ISL_495992\|2020-05-2 | ···················· | ···················· | ····················· | ···················· | ···················· | ···················· |
| F | 8224 | hCoV-19/USA/WA-S1567/2020\|EPI_ISL_495993\|2020-05-2 | ···················· | ···················· | ····················· | ···················· | ···················· | ···················· |
| F | 8225 | hCoV-19/USA/WA-S1569/2020\|EPI_ISL_495995\|2020-05-2 | ···················· | ···················· | ····················· | ···················· | ···················· | ···················· |
| F | 8226 | hCoV-19/USA/WA-S1570/2020\|EPI_ISL_495996\|2020-05-2 | ···················· | ···················· | ····················· | ···················· | ···················· | ···················· |
| F | 8227 | hCoV-19/USA/WA-S1571/2020\|EPI_ISL_495997\|2020-05-2 | ···················· | ···················· | ····················· | ···················· | ···················· | ···················· |
| F | 8228 | hCoV-19/USA/WA-S1572/2020\|EPI_ISL_495998\|2020-05-2 | ···················· | ···················· | ····················· | ···················· | ···················· | ···················· |
| F | 8229 | hCoV-19/USA/WA-S1573/2020\|EPI_ISL_495999\|2020-05-2 | ···················· | ···················· | ····················· | ···················· | ···················· | ···················· |
| F | 8230 | hCoV-19/Senegal/9068/2020\|EPI_ISL_498236\|2020-04-2 | ···················· | ···················· | ····················· | ···················· | ···················· | ···················· |
| F | 8231 | hCoV-19/Senegal/45436/2020\|EPI_ISL_498238\|2020-05- | ···················· | ···················· | ····················· | ···················· | ···················· | ···················· |
| F | 8232 | hCoV-19/Senegal/55499/2020\|EPI_ISL_498242\|2020-06- | ···················· | ···················· | ····················· | ···················· | ···················· | ···················· |
| F | 8233 | hCoV-19/Senegal/57273/2020\|EPI_ISL_498243\|2020-06- | ···················· | ···················· | ····················· | ···················· | ···················· | ···················· |
| F | 8234 | hCoV-19/Senegal/64716/2020\|EPI_ISL_498250\|2020-06- | ···················· | ···················· | ····················· | ···················· | ···················· | ···················· |
| F | 8235 | hCoV-19/Bangladesh/BCSIR-NILMRC-260/2020\|EPI_ISL_4 | ···················· | ···················· | ····················· | ···················· | ···················· | ···················· |
| F | 8237 | hCoV-19/USA/VA-DCLS-0693/2020\|EPI_ISL_500854\|2020- | ···················· | ···················· | ····················· | ···················· | ···················· | ···················· |
| F | 8238 | hCoV-19/USA/VA-DCLS-0694/2020\|EPI_ISL_500855\|2020- | ···················· | ···················· | ····················· | ···················· | ···················· | ···················· |
| F | 8239 | hCoV-19/USA/VA-DCLS-0700/2020\|EPI_ISL_500856\|2020- | ···················· | ···················· | ····················· | ···················· | ···················· | ···················· |
| F | 8240 | hCoV-19/USA/VA-DCLS-0701/2020\|EPI_ISL_500857\|2020- | ···················· | ···················· | ····················· | ···················· | ···················· | ···················· |
| F | 8241 | hCoV-19/USA/VA-DCLS-0702/2020\|EPI_ISL_500858\|2020- | ···················· | ···················· | ····················· | ···················· | ···················· | ···················· |
| F | 8242 | hCoV-19/USA/VA-DCLS-0705/2020\|EPI_ISL_500859\|2020- | ···················· | ···················· | ····················· | ···················· | ···················· | ···················· |
| F | 8243 | hCoV-19/USA/VA-DCLS-0707/2020\|EPI_ISL_500860\|2020- | ···················· | ···················· | ····················· | ···················· | ···················· | ···················· |
| F | 8244 | hCoV-19/USA/VA-DCLS-0708/2020\|EPI_ISL_500861\|2020- | ···················· | ···················· | ····················· | ···················· | ···················· | ···················· |
| F | 8245 | hCoV-19/USA/VA-DCLS-0709/2020\|EPI_ISL_500862\|2020- | ···················· | ···················· | ····················· | ···················· | ···················· | ···················· |
| F | 8246 | hCoV-19/USA/VA-DCLS-0711/2020\|EPI_ISL_500863\|2020- | ···················· | ···················· | ····················· | ···················· | ···················· | ···················· |
| F | 8247 | hCoV-19/Switzerland/BL-ETHZ-190086/2020\|EPI_ISL_50 | ···················· | ···················· | ····················· | ···················· | ···················· | ···················· |
| F | 8248 | hCoV-19/Switzerland/VD-ETHZ-190087/2020\|EPI_ISL_50 | ···················· | ···················· | ····················· | ···················· | ···················· | ···················· |
| F | 8249 | hCoV-19/Switzerland/BS-ETHZ-190088/2020\|EPI_ISL_50 | ···················· | ···················· | ····················· | ···················· | ···················· | ···················· |
| F | 8250 | hCoV-19/Switzerland/BL-ETHZ-190090/2020\|EPI_ISL_50 | ···················· | ···················· | ····················· | ···················· | ···················· | ···················· |
| F | 8251 | hCoV-19/India/GJ-GBRC288/2020\|EPI_ISL_500946\|2020- | ···················· | ···················· | ····················· | ···················· | ···················· | ···················· |
| F | 8252 | hCoV-19/India/GJ-GBRC317/2020\|EPI_ISL_500947\|2020- | ···················· | ···················· | ····················· | ···················· | ···················· | ···················· |
| F | 8253 | hCoV-19/India/GJ-GBRC321/2020\|EPI_ISL_500948\|2020- | ···················· | ···················· | ····················· | ···················· | ···················· | ···················· |
| F | 8254 | hCoV-19/India/GJ-GBRC322a/2020\|EPI_ISL_500949\|2020 | ···················· | ···················· | ····················· | ···················· | ···················· | ···················· |
| F | 8255 | hCoV-19/India/GJ-GBRC322b/2020\|EPI_ISL_500950\|2020 | ···················· | ···················· | ····················· | ···················· | ···················· | ···················· |
| F | 8256 | hCoV-19/England/201790215/2020\|EPI_ISL_500952\|2020 | ···················· | ···················· | ····················· | ···················· | ···················· | ···················· |
| F | 8257 | hCoV-19/England/201881261/2020\|EPI_ISL_500953\|2020 | ···················· | ···················· | ····················· | ···················· | ···················· | ···················· |
| F | 8258 | hCoV-19/Northern Ireland/NIRE-106C50/2020\|EPI_ISL_ | ···················· | ···················· | ····················· | ···················· | ···················· | ···················· |
| F | 8259 | hCoV-19/Northern Ireland/NIRE-106A56/2020\|EPI_ISL_ | ···················· | ···················· | ····················· | ···················· | ···················· | ···················· |
| F | 8260 | hCoV-19/Northern Ireland/NIRE-106E96/2020\|EPI_ISL_ | ···················· | ···················· | ····················· | ···················· | ···················· | ···················· |
| F | 8261 | hCoV-19/Northern Ireland/NIRE-106B26/2020\|EPI_ISL_ | ···················· | ···················· | ····················· | ···················· | ···················· | ···················· |
| F | 8262 | hCoV-19/Northern Ireland/NIRE-106AB0/2020\|EPI_ISL_ | ···················· | ···················· | ····················· | ···················· | ···················· | ···················· |
| F | 8263 | hCoV-19/Northern Ireland/NIRE-1069FF/2020\|EPI_ISL_ | ···················· | ···················· | ····················· | ···················· | ···················· | ···················· |
| F | 8264 | hCoV-19/Northern Ireland/NIRE-1066AD/2020\|EPI_ISL_ | ···················· | ···················· | ····················· | ···················· | ···················· | ···················· |
| F | 8265 | hCoV-19/Northern Ireland/NIRE-10650A/2020\|EPI_ISL_ | ···················· | ···················· | ····················· | ···················· | ···················· | ···················· |
| F | 8266 | hCoV-19/Northern Ireland/NIRE-106F0C/2020\|EPI_ISL_ | ···················· | ···················· | ····················· | ···················· | ···················· | ···················· |
| F | 8267 | hCoV-19/South Africa/KRISP-K000607/2020\|EPI_ISL_50 | ···················· | ···················· | ····················· | ···················· | ··T················· | ···················· |
| F | 8268 | hCoV-19/South Africa/KRISP-K000608/2020\|EPI_ISL_50 | ···················· | ···················· | ····················· | ···················· | ···················· | ···················· |
| F | 8269 | hCoV-19/South Africa/KRISP-K000609/2020\|EPI_ISL_50 | ···················· | ···················· | ····················· | ···················· | ···················· | ···················· |
| F | 8270 | hCoV-19/South Africa/KRISP-K000611/2020\|EPI_ISL_50 | ···················· | ···················· | ····················· | ···················· | ···················· | ···················· |
| F | 8271 | hCoV-19/South Africa/KRISP-K000612/2020\|EPI_ISL_50 | ···················· | ···················· | ····················· | ···················· | ···················· | ···················· |
| F | 8272 | hCoV-19/South Africa/KRISP-K000613/2020\|EPI_ISL_50 | ···················· | ···················· | ····················· | ···················· | ···················· | ···················· |
| F | 8273 | hCoV-19/South Africa/KRISP-K000614/2020\|EPI_ISL_50 | ···················· | ···················· | ····················· | ···················· | ···················· | ···················· |
| F | 8274 | hCoV-19/South Africa/KRISP-K000615/2020\|EPI_ISL_50 | ···················· | ···················· | ····················· | ···················· | ···················· | ···················· |
| F | 8275 | hCoV-19/South Africa/KRISP-K000617/2020\|EPI_ISL_50 | ···················· | ···················· | ····················· | ···················· | ···················· | ···················· |
| F | 8276 | hCoV-19/South Africa/KRISP-K000618/2020\|EPI_ISL_50 | ···················· | ···················· | ····················· | ···················· | ···················· | ···················· |
| F | 8277 | hCoV-19/South Africa/KRISP-K000619/2020\|EPI_ISL_50 | ···················· | ···················· | ····················· | ···················· | ···················· | ···················· |
| F | 8278 | hCoV-19/South Africa/KRISP-K000622/2020\|EPI_ISL_50 | ···················· | ···················· | ····················· | ···················· | ···················· | ···················· |
| F | 8279 | hCoV-19/South Africa/KRISP-K000623/2020\|EPI_ISL_50 | ···················· | ···················· | ····················· | ···················· | ···················· | ···················· |
| F | 8280 | hCoV-19/South Africa/KRISP-K000624/2020\|EPI_ISL_50 | ···················· | ···················· | ····················· | ···················· | ···················· | ···················· |
| F | 8281 | hCoV-19/South Africa/KRISP-K000628/2020\|EPI_ISL_50 | ···················· | ···················· | ····················· | ···················· | ···················· | ···················· |
| F | 8282 | hCoV-19/South Africa/KRISP-K000630/2020\|EPI_ISL_50 | ···················· | ···················· | ····················· | ···················· | ···················· | ···················· |
| F | 8283 | hCoV-19/South Africa/KRISP-K000632/2020\|EPI_ISL_50 | ···················· | ···················· | ····················· | ···················· | ···················· | ···················· |
| F | 8284 | hCoV-19/South Africa/KRISP-K000633/2020\|EPI_ISL_50 | ···················· | ···················· | ····················· | ···················· | ···················· | ···················· |
| F | 8285 | hCoV-19/South Africa/KRISP-K000635/2020\|EPI_ISL_50 | ···················· | ···················· | ····················· | ···················· | ···················· | ···················· |
| F | 8286 | hCoV-19/South Africa/KRISP-K000636/2020\|EPI_ISL_50 | ···················· | ···················· | ····················· | ···················· | ···················· | ···················· |
| F | 8287 | hCoV-19/South Africa/KRISP-K000637/2020\|EPI_ISL_50 | ···················· | ···················· | ····················· | ···················· | ···················· | ···················· |
| F | 8288 | hCoV-19/South Africa/KRISP-K000639/2020\|EPI_ISL_50 | ···················· | ···················· | ····················· | ···················· | ···················· | ···················· |
| F | 8289 | hCoV-19/South Africa/KRISP-K000640/2020\|EPI_ISL_50 | ···················· | ···················· | ····················· | ···················· | ···················· | ···················· |
| F | 8290 | hCoV-19/South Africa/KRISP-K000641/2020\|EPI_ISL_50 | ···················· | ···················· | ····················· | ···················· | ···················· | ···················· |

| | | | | | | | | |
|---|---|---|---|---|---|---|---|---|
| F | 8291 hCoV-19/South Africa/KRISP-K000643/2020\|EPI_ISL_50 | ···················· | ···················· | ···················· | ···················· | ···················· | ···················· | |
| F | 8292 hCoV-19/South Africa/KRISP-K000649/2020\|EPI_ISL_50 | ···················· | ···················· | ···················· | ···················· | ···················· | ···················· | |
| F | 8293 hCoV-19/South Africa/KRISP-K000650/2020\|EPI_ISL_50 | ···················· | ···················· | ···················· | ···················· | ···················· | ···················· | |
| F | 8294 hCoV-19/South Africa/KRISP-K000651/2020\|EPI_ISL_50 | ···················· | ···················· | ···················· | ···················· | ···················· | ···················· | |
| F | 8295 hCoV-19/South Africa/KRISP-K000652/2020\|EPI_ISL_50 | ···················· | ···················· | ···················· | ···················· | ···················· | ···················· | |
| F | 8296 hCoV-19/South Africa/KRISP-K000654/2020\|EPI_ISL_50 | ···················· | ···················· | ···················· | ···················· | ···················· | ···················· | |
| F | 8297 hCoV-19/South Africa/KRISP-K000658/2020\|EPI_ISL_50 | ···················· | ···················· | ···················· | ···················· | ···················· | ···················· | |
| F | 8298 hCoV-19/Spain/CT-IBV-003151/2020\|EPI_ISL_510248\|20 | ···················· | ···················· | ···················· | ···················· | ···················· | ···················· | |
| F | 8302 hCoV-19/Spain/CT-IBV-003155/2020\|EPI_ISL_510252\|20 | ···················· | ···················· | ···················· | ···················· | ···················· | ···················· | |
| F | 8303 hCoV-19/Spain/CT-IBV-003156/2020\|EPI_ISL_510253\|20 | ···················· | ···················· | ···················· | ···················· | ···················· | ···················· | |
| F | 8304 hCoV-19/Spain/CT-IBV-003160/2020\|EPI_ISL_510256\|20 | ···················· | ···················· | ···················· | ···················· | ···················· | ···················· | |
| F | 8305 hCoV-19/Spain/CT-IBV-003161/2020\|EPI_ISL_510257\|20 | ···················· | ···················· | ···················· | ···················· | ···················· | ···················· | |
| F | 8307 hCoV-19/Spain/CT-IBV-003163/2020\|EPI_ISL_510259\|20 | ···················· | ···················· | ···················· | ···················· | ···················· | ···················· | |
| F | 8308 hCoV-19/Spain/CT-IBV-003164/2020\|EPI_ISL_510260\|20 | ···················· | ···················· | ···················· | ···················· | ···················· | ···················· | |
| F | 8311 hCoV-19/Spain/CT-IBV-003167/2020\|EPI_ISL_510263\|20 | ···················· | ···················· | ···················· | ···················· | ···················· | ···················· | |
| F | 8313 hCoV-19/Spain/CT-IBV-003176/2020\|EPI_ISL_510265\|20 | ···················· | ···················· | ···················· | ···················· | ···················· | ···················· | |
| F | 8315 hCoV-19/Spain/GA-IBV-004025/2020\|EPI_ISL_510268\|20 | ···················· | ···················· | ···················· | ···················· | ···················· | ···················· | |
| F | 8316 hCoV-19/Spain/GA-IBV-004027/2020\|EPI_ISL_510269\|20 | ···················· | ···················· | ···················· | ···················· | ···················· | ···················· | |
| F | 8317 hCoV-19/Spain/GA-IBV-004029/2020\|EPI_ISL_510271\|20 | ···················· | ···················· | ···················· | ···················· | ···················· | ···················· | |
| F | 8319 hCoV-19/Spain/GA-IBV-004032/2020\|EPI_ISL_510273\|20 | ···················· | ···················· | ···················· | ···················· | ···················· | ···················· | |
| F | 8320 hCoV-19/Spain/GA-IBV-004035/2020\|EPI_ISL_510274\|20 | ···················· | ···················· | ···················· | ···················· | ···················· | ···················· | |
| F | 8321 hCoV-19/Spain/GA-IBV-004036/2020\|EPI_ISL_510275\|20 | ···················· | ···················· | ···················· | ···················· | ···················· | ···················· | |
| F | 8326 hCoV-19/Spain/GA-IBV-004044/2020\|EPI_ISL_510283\|20 | ···················· | ···················· | ···················· | ···················· | ···················· | ···················· | |
| F | 8327 hCoV-19/Spain/GA-IBV-004045/2020\|EPI_ISL_510284\|20 | ···················· | ···················· | ···················· | ···················· | ···················· | ···················· | |
| F | 8331 hCoV-19/Spain/GA-IBV-004051/2020\|EPI_ISL_510290\|20 | ···················· | ···················· | ···················· | ···················· | ···················· | ···················· | |
| F | 8332 hCoV-19/Spain/GA-IBV-004052/2020\|EPI_ISL_510291\|20 | ···················· | ···················· | ···················· | ···················· | ·NNN················ | ···················· | 1 |
| F | 8334 hCoV-19/Spain/GA-IBV-004054/2020\|EPI_ISL_510293\|20 | ···················· | ···················· | ···················· | ···················· | ···················· | ···················· | |
| F | 8335 hCoV-19/Spain/GA-IBV-004055/2020\|EPI_ISL_510294\|20 | ···················· | ···················· | ···················· | ···················· | ···················· | ···················· | |
| F | 8338 hCoV-19/Spain/GA-IBV-004060/2020\|EPI_ISL_510298\|20 | ···················· | ···················· | ···················· | ···················· | ···················· | ···················· | |
| F | 8339 hCoV-19/Portugal/PT0950/2020\|EPI_ISL_511500\|2020-0 | ···················· | ···················· | ···················· | ···················· | ···················· | ···················· | |
| F | 8340 hCoV-19/Portugal/PT0951/2020\|EPI_ISL_511501\|2020-0 | ···················· | ···················· | ···················· | ···················· | ···················· | ···················· | |
| F | 8341 hCoV-19/Portugal/PT1037/2020\|EPI_ISL_511584\|2020-0 | ···················· | ···················· | ···················· | ···················· | ···················· | ···················· | |
| F | 8342 hCoV-19/Portugal/PT1047/2020\|EPI_ISL_511594\|2020-0 | ···················· | ···················· | ···················· | ···················· | ···················· | ···················· | |
| F | 8343 hCoV-19/Portugal/PT1051/2020\|EPI_ISL_511598\|2020-0 | ···················· | ···················· | ···················· | ···················· | ···················· | ···················· | |
| F | 8344 hCoV-19/Portugal/PT1053/2020\|EPI_ISL_511600\|2020-0 | ···················· | ···················· | ···················· | ···················· | ···················· | ···················· | |
| F | 8345 hCoV-19/Portugal/PT1056/2020\|EPI_ISL_511603\|2020-0 | ···················· | ···················· | ···················· | ···················· | ···················· | ···················· | |
| F | 8346 hCoV-19/Portugal/PT1064/2020\|EPI_ISL_511611\|2020-0 | ···················· | ···················· | ···················· | ···················· | ···················· | ···················· | |
| F | 8347 hCoV-19/Singapore/489ilmn/2020\|EPI_ISL_512822\|2020 | ···················· | ···················· | ···················· | ···················· | ···················· | ···················· | |
| F | 8348 hCoV-19/Singapore/787/2020\|EPI_ISL_512823\|2020-04- | ···················· | ···················· | ···················· | ···················· | ···················· | ···················· | |
| F | 8349 hCoV-19/Singapore/788/2020\|EPI_ISL_512824\|2020-04- | ···················· | ···················· | ···················· | ···················· | ···················· | ···················· | |
| F | 8350 hCoV-19/Singapore/789/2020\|EPI_ISL_512825\|2020-04- | ···················· | ···················· | ···················· | ···················· | ···················· | ···················· | |
| F | 8351 hCoV-19/Singapore/790/2020\|EPI_ISL_512826\|2020-04- | ···················· | ···················· | ···················· | ···················· | ···················· | ···················· | |
| F | 8352 hCoV-19/Singapore/791/2020\|EPI_ISL_512827\|2020-06- | ···················· | ···················· | ···················· | ···················· | ···················· | ···················· | |
| F | 8353 hCoV-19/Singapore/792/2020\|EPI_ISL_512828\|2020-06- | ···················· | ···················· | ···················· | ···················· | ···················· | ···················· | |
| F | 8354 hCoV-19/Singapore/793/2020\|EPI_ISL_512829\|2020-06- | ···················· | ···················· | ···················· | ···················· | ···················· | ···················· | |
| F | 8355 hCoV-19/Singapore/794/2020\|EPI_ISL_512830\|2020-04- | ···················· | ···················· | ···················· | ···················· | ···················· | ···················· | |
| F | 8356 hCoV-19/Singapore/796/2020\|EPI_ISL_512832\|2020-07- | ···················· | ···················· | ···················· | ···················· | ···················· | ···················· | |
| F | 8357 hCoV-19/SaudiArabia/KAUST-JEDDAH322/2020\|EPI_ISL_5 | ···················· | ···················· | ···················· | ···················· | ···················· | ···················· | |
| F | 8358 hCoV-19/SaudiArabia/KAUST-JEDDAH323/2020\|EPI_ISL_5 | ···················· | ·A·················· | ···················· | ···················· | ···················· | ··A················· | |
| F | 8359 hCoV-19/SaudiArabia/KAUST-JEDDAH324/2020\|EPI_ISL_5 | ···················· | ···················· | ···················· | ···················· | ···················· | ···················· | |
| F | 8360 hCoV-19/SaudiArabia/KAUST-JEDDAH329/2020\|EPI_ISL_5 | ···················· | ···················· | ···················· | ···················· | ······NNN··········· | ···················· | 1 |
| F | 8361 hCoV-19/SaudiArabia/KAUST-JEDDAH330/2020\|EPI_ISL_5 | ···················· | ···················· | ···················· | ···················· | ···················· | ···················· | |
| F | 8362 hCoV-19/SaudiArabia/KAUST-JEDDAH331/2020\|EPI_ISL_5 | ···················· | ···················· | ···················· | ···················· | ···················· | ···················· | |
| F | 8363 hCoV-19/SaudiArabia/KAUST-JEDDAH332/2020\|EPI_ISL_5 | ···················· | ···················· | ···················· | ···················· | ···················· | ···················· | |
| F | 8364 hCoV-19/SaudiArabia/KAUST-JEDDAH466/2020\|EPI_ISL_5 | ···················· | ···················· | ···················· | ···················· | ···················· | ···················· | |
| F | 8365 hCoV-19/SaudiArabia/KAUST-JEDDAH467/2020\|EPI_ISL_5 | ···················· | ···················· | ···················· | ···················· | ···················· | ···················· | |
| F | 8366 hCoV-19/SaudiArabia/KAUST-JEDDAH468/2020\|EPI_ISL_5 | ···················· | ···················· | ···················· | ···················· | ···················· | ···················· | |
| F | 8367 hCoV-19/SaudiArabia/KAUST-JEDDAH469/2020\|EPI_ISL_5 | ···················· | ···················· | ···················· | ···················· | ···················· | ···················· | |
| F | 8368 hCoV-19/SaudiArabia/KAUST-JEDDAH471/2020\|EPI_ISL_5 | ···················· | ···················· | ···················· | ···················· | ···················· | ···················· | |
| F | 8369 hCoV-19/SaudiArabia/KAUST-JEDDAH473/2020\|EPI_ISL_5 | ···················· | ···················· | ···················· | ···················· | ···················· | ···················· | |
| F | 8370 hCoV-19/SaudiArabia/KAUST-JEDDAH479/2020\|EPI_ISL_5 | ···················· | ···················· | ···················· | ···················· | ···················· | ···················· | |
| F | 8371 hCoV-19/SaudiArabia/KAUST-JEDDAH482/2020\|EPI_ISL_5 | ···················· | ···················· | ···················· | ···················· | ···················· | ···················· | |
| F | 8372 hCoV-19/SaudiArabia/KAUST-JEDDAH483/2020\|EPI_ISL_5 | ···················· | ···················· | ···················· | ···················· | ···················· | ···················· | |
| F | 8373 hCoV-19/SaudiArabia/KAUST-JEDDAH486/2020\|EPI_ISL_5 | ···················· | ···················· | ···················· | ···················· | ···················· | ···················· | |
| F | 8374 hCoV-19/SaudiArabia/KAUST-JEDDAH487/2020\|EPI_ISL_5 | ···················· | ···················· | ···················· | ···················· | ···················· | ···················· | |
| F | 8375 hCoV-19/SaudiArabia/KAUST-JEDDAH488/2020\|EPI_ISL_5 | ···················· | ···················· | ···················· | ···················· | ···················· | ···················· | |
| F | 8376 hCoV-19/SaudiArabia/KAUST-JEDDAH490/2020\|EPI_ISL_5 | ···················· | ···················· | ···················· | ···················· | ···················· | ···················· | |
| F | 8377 hCoV-19/SaudiArabia/KAUST-JEDDAH492/2020\|EPI_ISL_5 | ···················· | ···················· | ···················· | ···················· | ···················· | ···················· | |
| F | 8378 hCoV-19/SaudiArabia/KAUST-JEDDAH493/2020\|EPI_ISL_5 | ···················· | ···················· | ···················· | ···················· | ···················· | ···················· | |
| F | 8379 hCoV-19/SaudiArabia/KAUST-JEDDAH494/2020\|EPI_ISL_5 | ···················· | ···················· | ···················· | ···················· | ···················· | ···················· | |
| F | 8382 hCoV-19/SaudiArabia/KAUST-JEDDAH504/2020\|EPI_ISL_5 | ···················· | ···················· | ···················· | ···················· | ···················· | ···················· | |
| F | 8383 hCoV-19/SaudiArabia/KAUST-JEDDAH505/2020\|EPI_ISL_5 | ···················· | ···················· | ···················· | ···················· | ···················· | ···················· | |
| F | 8391 hCoV-19/Latvia/104/2020\|EPI_ISL_515191\|2020-08-05 | ···················· | ···················· | ···················· | ···················· | ···················· | ···················· | |
| F | 8392 hCoV-19/USA/MI-MDHHS-SC21685/2020\|EPI_ISL_516412\|2 | ···················· | ···················· | ···················· | ···················· | ·NNN················ | ···················· | 1 |
| F | 8393 hCoV-19/North Macedonia/2499/2020\|EPI_ISL_516413\|2 | ···················· | ···················· | ···················· | ···················· | ···················· | ···················· | |
| F | 8394 hCoV-19/North Macedonia/2531/2020\|EPI_ISL_516414\|2 | ···················· | ···················· | ···················· | ···················· | ···················· | ···················· | |
| F | 8395 hCoV-19/North Macedonia/2819/2020\|EPI_ISL_516415\|2 | ···················· | ···················· | ···················· | ···················· | ···················· | ···················· | |
| F | 8396 hCoV-19/North Macedonia/2851/2020\|EPI_ISL_516416\|2 | ···················· | ···················· | ···················· | ···················· | ···················· | ···················· | |
| F | 8397 hCoV-19/North Macedonia/2856/2020\|EPI_ISL_516417\|2 | ···················· | ···················· | ···················· | ···················· | ···················· | ···················· | |
| F | 8398 hCoV-19/North Macedonia/2907/2020\|EPI_ISL_516418\|2 | ···················· | ····G··············· | ···················· | ···················· | ···················· | ···················· | |

| | | | | | | | |
|---|---|---|---|---|---|---|---|
| F | 8399 hCoV-19/North Macedonia/2979/2020\|EPI_ISL_516419\|2 | ···················· | ···················· | ···················· | ···················· | ···················· | ···················· |
| F | 8400 hCoV-19/North Macedonia/3017/2020\|EPI_ISL_516420\|2 | ···················· | ···················· | ···················· | ···················· | ···················· | ···················· |
| F | 8401 hCoV-19/North Macedonia/3122/2020\|EPI_ISL_516421\|2 | ···················· | ···················· | ···················· | ···················· | ···················· | ···················· |
| F | 8402 hCoV-19/North Macedonia/3193/2020\|EPI_ISL_516422\|2 | ···················· | ···················· | ···················· | ···················· | ···················· | ···················· |
| F | 8403 hCoV-19/North Macedonia/3310/2020\|EPI_ISL_516423\|2 | ···················· | ···················· | ···················· | ···················· | ···················· | ···················· |
| F | 8404 hCoV-19/North Macedonia/3861/2020\|EPI_ISL_516424\|2 | ···················· | ···················· | ···················· | ···················· | ···················· | ···················· |
| F | 8405 hCoV-19/North Macedonia/3990/2020\|EPI_ISL_516425\|2 | ···················· | ···················· | ···················· | ···················· | ···················· | ···················· |
| F | 8406 hCoV-19/North Macedonia/5016/2020\|EPI_ISL_516426\|2 | ···················· | ···················· | ···················· | ···················· | ···················· | ···················· |
| F | 8407 hCoV-19/North Macedonia/5220/2020\|EPI_ISL_516427\|2 | ···················· | ···················· | ···················· | ···················· | ···················· | ···················· |
| F | 8408 hCoV-19/North Macedonia/6497/2020\|EPI_ISL_516428\|2 | ···················· | ···················· | ···················· | ···················· | ···················· | ···················· |
| F | 8409 hCoV-19/North Macedonia/7229/2020\|EPI_ISL_516429\|2 | ···················· | ···················· | ···················· | ···················· | ···················· | ···················· |
| F | 8410 hCoV-19/North Macedonia/7245/2020\|EPI_ISL_516430\|2 | ···················· | ···················· | ···················· | ···················· | ···················· | ···················· |
| F | 8411 hCoV-19/North Macedonia/7808/2020\|EPI_ISL_516431\|2 | ···················· | ···················· | ···················· | ···················· | ···················· | ···················· |
| F | 8412 hCoV-19/USA/WI-UW-1006/2020\|EPI_ISL_516432\|2020-07 | ···················· | ···················· | ···················· | ···················· | ···················· | ···················· |
| F | 8413 hCoV-19/USA/WI-UW-1007/2020\|EPI_ISL_516433\|2020-07 | ···················· | ···················· | ···················· | ···················· | ···················· | ···················· |
| F | 8414 hCoV-19/USA/WI-UW-1008/2020\|EPI_ISL_516434\|2020-07 | ···················· | ···················· | ···················· | ···················· | ···················· | ···················· |
| F | 8415 hCoV-19/USA/IL-UW-1009/2020\|EPI_ISL_516435\|2020-07 | ···················· | ···················· | ···················· | ···················· | ···················· | ···················· |
| F | 8416 hCoV-19/USA/WI-UW-1010/2020\|EPI_ISL_516436\|2020-07 | ···················· | ·A·················· | ···················· | ···················· | ···················· | ···················· |
| F | 8417 hCoV-19/USA/WI-UW-1011/2020\|EPI_ISL_516437\|2020-07 | ···················· | ···················· | ···················· | ···················· | ···················· | ···················· |
| F | 8418 hCoV-19/USA/WI-UW-1012/2020\|EPI_ISL_516438\|2020-07 | ···················· | ···················· | ···················· | ···················· | ···················· | ···················· |
| F | 8419 hCoV-19/USA/WI-UW-1013/2020\|EPI_ISL_516439\|2020-07 | ···················· | ···················· | ···················· | ···················· | ···················· | ···················· |
| F | 8420 hCoV-19/USA/WI-UW-1014/2020\|EPI_ISL_516440\|2020-07 | ···················· | ···················· | ···················· | ···················· | ···················· | ···················· |
| F | 8421 hCoV-19/USA/WI-UW-1015/2020\|EPI_ISL_516441\|2020-07 | ···················· | ···················· | ···················· | ···················· | ···················· | ···················· |
| F | 8422 hCoV-19/USA/WI-UW-1016/2020\|EPI_ISL_516442\|2020-07 | ···················· | ···················· | ···················· | ···················· | ···················· | ···················· |
| F | 8423 hCoV-19/USA/WI-UW-1017/2020\|EPI_ISL_516443\|2020-07 | ···················· | ···················· | ···················· | ···················· | ···················· | ···················· |
| F | 8424 hCoV-19/USA/WI-UW-1018/2020\|EPI_ISL_516444\|2020-07 | ···················· | ···················· | ···················· | ···················· | ···················· | ···················· |
| F | 8425 hCoV-19/USA/WI-UW-1019/2020\|EPI_ISL_516445\|2020-07 | ···················· | ···················· | ···················· | ···················· | ···················· | ···················· |
| F | 8426 hCoV-19/USA/WI-UW-1020/2020\|EPI_ISL_516446\|2020-08 | ···················· | ···················· | ···················· | ···················· | ···················· | ···················· |
| F | 8427 hCoV-19/USA/WI-UW-1021/2020\|EPI_ISL_516447\|2020-07 | ···················· | ···················· | ···················· | ···················· | ···················· | ···················· |
| F | 8428 hCoV-19/USA/WI-UW-1022/2020\|EPI_ISL_516448\|2020-07 | ···················· | ···················· | ···················· | ···················· | ···················· | ···················· |
| F | 8430 hCoV-19/USA/WI-UW-1025/2020\|EPI_ISL_516451\|2020-08 | ···················· | ···················· | ···················· | ···················· | ···················· | ···················· |
| F | 8431 hCoV-19/USA/WI-UW-1026/2020\|EPI_ISL_516452\|2020-08 | ···················· | ···················· | ···················· | ···················· | ···················· | ···················· |
| F | 8432 hCoV-19/USA/WI-UW-1027/2020\|EPI_ISL_516453\|2020-08 | ···················· | ···················· | ···················· | ···················· | ···················· | ···················· |
| F | 8433 hCoV-19/USA/WI-UW-1028/2020\|EPI_ISL_516454\|2020-08 | ···················· | ···················· | ···················· | ···················· | ···················· | ···················· |
| F | 8434 hCoV-19/USA/WI-UW-1029/2020\|EPI_ISL_516455\|2020-08 | ···················· | ···················· | ···················· | ···················· | ···················· | ···················· |
| F | 8435 hCoV-19/USA/WI-UW-1030/2020\|EPI_ISL_516456\|2020-08 | ···················· | ···················· | ···················· | ···················· | ···················· | ···················· |
| F | 8436 hCoV-19/USA/WI-UW-1031/2020\|EPI_ISL_516457\|2020-07 | ···················· | ···················· | ···················· | ···················· | ···················· | ···················· |
| F | 8438 hCoV-19/USA/WI-UW-1033/2020\|EPI_ISL_516459\|2020-07 | ···················· | ···················· | ···················· | ···················· | ···················· | ···················· |
| F | 8439 hCoV-19/USA/WI-UW-1034/2020\|EPI_ISL_516460\|2020-08 | ···················· | ···················· | ···················· | ···················· | ···················· | ···················· |
| F | 8440 hCoV-19/USA/WI-UW-1035/2020\|EPI_ISL_516461\|2020-08 | ···················· | ···················· | ···················· | ···················· | ···················· | ···················· |
| F | 8441 hCoV-19/USA/WI-UW-1036/2020\|EPI_ISL_516462\|2020-08 | ···················· | ···················· | ···················· | ···················· | ···················· | ···················· |
| F | 8442 hCoV-19/USA/WI-UW-1037/2020\|EPI_ISL_516463\|2020-08 | ···················· | ···················· | ···················· | ···················· | ···················· | ···················· |
| F | 8443 hCoV-19/USA/WI-UW-1038/2020\|EPI_ISL_516464\|2020-07 | ···················· | ···················· | ···················· | ···················· | ···················· | ···················· |
| F | 8445 hCoV-19/USA/WI-UW-1040/2020\|EPI_ISL_516466\|2020-07 | ···················· | ···················· | ···················· | ···················· | ···················· | ···················· |
| F | 8446 hCoV-19/USA/WI-UW-1041/2020\|EPI_ISL_516467\|2020-08 | ···················· | ···················· | ···················· | ···················· | ···················· | ···················· |
| F | 8447 hCoV-19/USA/WI-UW-1042/2020\|EPI_ISL_516468\|2020-08 | ···················· | ···················· | ···················· | ···················· | ···················· | ···················· |
| F | 8448 hCoV-19/USA/WI-UW-1043/2020\|EPI_ISL_516469\|2020-08 | ···················· | ···················· | ···················· | ···················· | ···················· | ···················· |
| F | 8449 hCoV-19/USA/WA-S1492/2020\|EPI_ISL_495918\|2020-05-2 | ···················· | ···················· | ···················· | ···················· | ···················· | ···················· |
| F | 8450 hCoV-19/USA/WA-S1493/2020\|EPI_ISL_495919\|2020-05-2 | ···················· | ···················· | ···················· | ···················· | ···················· | ···················· |
| F | 8451 hCoV-19/USA/WA-S1494/2020\|EPI_ISL_495920\|2020-05-2 | ···················· | ···················· | ···················· | ···················· | ···················· | ···················· |
| F | 8452 hCoV-19/USA/WA-S1495/2020\|EPI_ISL_495921\|2020-05-2 | ···················· | ···················· | ···················· | ···················· | ···················· | ···················· |
| F | 8453 hCoV-19/USA/WA-S1496/2020\|EPI_ISL_495922\|2020-05-2 | ···················· | ···················· | ···················· | ···················· | ···················· | ···················· |
| F | 8454 hCoV-19/USA/WA-S1497/2020\|EPI_ISL_495923\|2020-05-2 | ···················· | ···················· | ···················· | ···················· | ···················· | ···················· |
| F | 8455 hCoV-19/USA/WA-S1498/2020\|EPI_ISL_495924\|2020-05-2 | ···················· | ···················· | ···················· | ···················· | ···················· | ···················· |
| F | 8456 hCoV-19/USA/WA-S1500/2020\|EPI_ISL_495926\|2020-05-2 | ···················· | ···················· | ···················· | ···················· | ···················· | ···················· |
| F | 8457 hCoV-19/USA/WA-S1501/2020\|EPI_ISL_495927\|2020-05-2 | ···················· | ···················· | ···················· | ···················· | ···················· | ···················· |
| F | 8458 hCoV-19/USA/WA-S1502/2020\|EPI_ISL_495928\|2020-05-2 | ···················· | ···················· | ···················· | ···················· | ···················· | ···················· |
| F | 8459 hCoV-19/USA/WA-S1503/2020\|EPI_ISL_495929\|2020-05-2 | ···················· | ···················· | ···················· | ···················· | ···················· | ···················· |
| F | 8460 hCoV-19/USA/WA-S1504/2020\|EPI_ISL_495930\|2020-05-2 | ···················· | ···················· | ···················· | ···················· | ···················· | ···················· |
| F | 8461 hCoV-19/USA/WA-S1505/2020\|EPI_ISL_495931\|2020-05-2 | ···················· | ···················· | ···················· | ···················· | ···················· | ···················· |
| F | 8462 hCoV-19/USA/WA-S1506/2020\|EPI_ISL_495932\|2020-05-2 | ···················· | ···················· | ···················· | ···················· | ···················· | ···················· |
| F | 8463 hCoV-19/USA/WA-S1508/2020\|EPI_ISL_495934\|2020-05-2 | ···················· | ···················· | ···················· | ···················· | ···················· | ···················· |
| F | 8464 hCoV-19/USA/WA-S1509/2020\|EPI_ISL_495935\|2020-05-2 | ···················· | ···················· | ···················· | ···················· | ···················· | ···················· |
| F | 8465 hCoV-19/USA/WA-S1510/2020\|EPI_ISL_495936\|2020-05-2 | ···················· | ···················· | ···················· | ···················· | ···················· | ···················· |
| F | 8466 hCoV-19/USA/WA-S1511/2020\|EPI_ISL_495937\|2020-05-2 | ···················· | ···················· | ···················· | ···················· | ···················· | ···················· |
| F | 8467 hCoV-19/USA/WA-S1512/2020\|EPI_ISL_495938\|2020-05-2 | ···················· | ···················· | ···················· | ···················· | ···················· | ···················· |
| F | 8468 hCoV-19/USA/WA-S1536/2020\|EPI_ISL_495962\|2020-05-2 | ···················· | ···················· | ···················· | ···················· | ···················· | ···················· |
| F | 8469 hCoV-19/USA/WA-S1537/2020\|EPI_ISL_495963\|2020-05-2 | ···················· | ···················· | ···················· | ···················· | ···················· | ···················· |
| F | 8470 hCoV-19/USA/WA-S1538/2020\|EPI_ISL_495964\|2020-05-2 | ···················· | ···················· | ···················· | ···················· | ···················· | ···················· |
| F | 8471 hCoV-19/USA/WA-S1539/2020\|EPI_ISL_495965\|2020-05-2 | ···················· | ···················· | ···················· | ···················· | ···················· | ···················· |
| F | 8472 hCoV-19/USA/WA-S1540/2020\|EPI_ISL_495966\|2020-05-2 | ···················· | ···················· | ···················· | ···················· | ···················· | ···················· |
| F | 8473 hCoV-19/USA/WA-S1541/2020\|EPI_ISL_495967\|2020-05-2 | ···················· | ···················· | ···················· | ···················· | ···················· | ···················· |
| F | 8474 hCoV-19/USA/WA-S1542/2020\|EPI_ISL_495968\|2020-05-2 | ···················· | ···················· | ···················· | ···················· | ···················· | ···················· |
| F | 8475 hCoV-19/USA/WA-S1552/2020\|EPI_ISL_495978\|2020-05-2 | ···················· | ···················· | ···················· | ···················· | ···················· | ···················· |
| F | 8476 hCoV-19/USA/WA-S1553/2020\|EPI_ISL_495979\|2020-05-2 | ···················· | ···················· | ···················· | ···················· | ···················· | ···················· |
| F | 8477 hCoV-19/USA/WA-S1554/2020\|EPI_ISL_495980\|2020-05-2 | ···················· | ···················· | ···················· | ···················· | ···················· | ···················· |
| F | 8478 hCoV-19/Vietnam/HCMC-05009/2020\|EPI_ISL_498180\|202 | ···················· | ···················· | ···················· | ···················· | ···················· | ···················· |
| F | 8479 hCoV-19/Vietnam/HCMC-35001/2020\|EPI_ISL_498181\|202 | ···················· | ···················· | ···················· | ···················· | ···················· | ···················· |
| F | 8480 hCoV-19/Vietnam/HCMC-35003/2020\|EPI_ISL_498182\|202 | ···················· | ···················· | ···················· | ···················· | ···················· | ···················· |
| F | 8481 hCoV-19/Vietnam/HCMC-35006/2020\|EPI_ISL_498183\|202 | ···················· | ···················· | ···················· | ···················· | ···················· | ···················· |

| | | | | | | | | |
|---|---|---|---|---|---|---|---|---|
| F | 8482 | hCoV-19/Vietnam/HCMC-35008/2020\|EPI_ISL_498184\|202 | ···················· | ···················· | ····················· | ···················· | ···················· | ···················· |
| F | 8483 | hCoV-19/Vietnam/HCMC-35009/2020\|EPI_ISL_498185\|202 | ···················· | ···················· | ····················· | ···················· | ···················· | ···················· |
| F | 8484 | hCoV-19/Vietnam/HCMC-35010/2020\|EPI_ISL_498186\|202 | ···················· | ···················· | ····················· | ···················· | ···················· | ···················· |
| F | 8485 | hCoV-19/Vietnam/HCMC-35012/2020\|EPI_ISL_498187\|202 | ···················· | ····A··············· | ····················· | ···················· | ···················· | ···················· |
| F | 8486 | hCoV-19/Vietnam/HCMC-35013/2020\|EPI_ISL_498188\|202 | ···················· | ····A··············· | ····················· | ···················· | ···················· | ···················· |
| F | 8487 | hCoV-19/Vietnam/HCMC-35014/2020\|EPI_ISL_498189\|202 | ···················· | ···················· | ····················· | ···················· | ···················· | ···················· |
| F | 8488 | hCoV-19/Vietnam/HCMC-35023/2020\|EPI_ISL_498190\|202 | ···················· | ···················· | ····················· | ···················· | ···················· | ···················· |
| F | 8489 | hCoV-19/Vietnam/HCMC-35030/2020\|EPI_ISL_498191\|202 | ···················· | ···················· | ····················· | ···················· | ···················· | ···················· |
| F | 8490 | hCoV-19/Vietnam/HCMC-35031/2020\|EPI_ISL_498192\|202 | ···················· | ···················· | ····················· | ···················· | ···················· | ···················· |
| F | 8491 | hCoV-19/England/NOTT-112D7C/2020\|EPI_ISL_499266\|20 | ···················· | ···················· | ····················· | ···················· | ···················· | ···················· |
| F | 8492 | hCoV-19/England/NOTT-112D8B/2020\|EPI_ISL_499267\|20 | ···················· | ···················· | ····················· | ···················· | ···················· | ···················· |
| F | 8493 | hCoV-19/England/NOTT-112DA9/2020\|EPI_ISL_499269\|20 | ···················· | ···················· | ····················· | ···················· | ···················· | ···················· |
| F | 8494 | hCoV-19/Norway/2846/2020\|EPI_ISL_500769\|2020-07-03 | ···················· | ···················· | ····················· | ···················· | ···················· | ···················· |
| F | 8495 | hCoV-19/Norway/2847/2020\|EPI_ISL_500770\|2020-07-03 | ···················· | ·A·················· | ····················· | ···················· | ···················· | ···················· |
| F | 8496 | hCoV-19/Norway/2848/2020\|EPI_ISL_500771\|2020-07-03 | ···················· | ·A·················· | ····················· | ···················· | ···················· | ···················· |
| F | 8497 | hCoV-19/Norway/2850/2020\|EPI_ISL_500773\|2020-07-02 | ···················· | ···················· | ····················· | ···················· | ···················· | ···················· |
| F | 8498 | hCoV-19/Norway/2851/2020\|EPI_ISL_500774\|2020-07-02 | ···················· | ·A·················· | ····················· | ···················· | ···················· | ···················· |
| F | 8499 | hCoV-19/Norway/2852/2020\|EPI_ISL_500775\|2020-07-03 | ···················· | ···················· | ····················· | ···················· | ···················· | ···················· |
| F | 8500 | hCoV-19/Norway/2853/2020\|EPI_ISL_500776\|2020-06-29 | ···················· | ···················· | ····················· | ···················· | ···················· | ···················· |
| F | 8502 | hCoV-19/Norway/2859/2020\|EPI_ISL_500782\|2020-07-04 | ···················· | ·A·················· | ····················· | ···················· | ···················· | ···················· |
| F | 8503 | hCoV-19/Norway/2860/2020\|EPI_ISL_500783\|2020-07-03 | ···················· | ···················· | ····················· | ···················· | ···················· | ···················· |
| F | 8504 | hCoV-19/Norway/2862/2020\|EPI_ISL_500785\|2020-07-06 | ···················· | ·A·················· | ····················· | ···················· | ···················· | ···················· |
| F | 8505 | hCoV-19/Norway/2864/2020\|EPI_ISL_500786\|2020-07-05 | ···················· | ···················· | ····················· | ···················· | ···················· | ···················· |
| F | 8506 | hCoV-19/Norway/2865/2020\|EPI_ISL_500787\|2020-07-07 | ···················· | ·A·················· | ····················· | ···················· | ···················· | ···················· |
| F | 8507 | hCoV-19/Norway/2870/2020\|EPI_ISL_500789\|2020-07-09 | ···················· | ···················· | ····················· | ···················· | ···················· | ···················· |
| F | 8508 | hCoV-19/Norway/2871/2020\|EPI_ISL_500790\|2020-07-08 | ···················· | ·A·················· | ····················· | ···················· | ···················· | ···················· |
| F | 8509 | hCoV-19/Norway/2872/2020\|EPI_ISL_500791\|2020-07-10 | ···················· | ·A·················· | ····················· | ···················· | ···················· | ···················· |
| F | 8510 | hCoV-19/Norway/2873/2020\|EPI_ISL_500792\|2020-07-10 | ···················· | ·A·················· | ····················· | ···················· | ···················· | ···················· |
| F | 8511 | hCoV-19/Norway/2875/2020\|EPI_ISL_500794\|2020-07-10 | ···················· | ···················· | ····················· | ···················· | ···················· | ···················· |
| F | 8512 | hCoV-19/Norway/2876/2020\|EPI_ISL_500795\|2020-07-08 | ···················· | ·A·················· | ····················· | ···················· | ···················· | ···················· |
| F | 8513 | hCoV-19/Norway/2877/2020\|EPI_ISL_500796\|2020-07-11 | ···················· | ···················· | ····················· | ···················· | ···················· | ···················· |
| F | 8514 | hCoV-19/Norway/2879/2020\|EPI_ISL_500797\|2020-07-13 | ···················· | ···················· | ····················· | ···················· | ···················· | ···················· |
| F | 8515 | hCoV-19/Norway/2881/2020\|EPI_ISL_500799\|2020-07-13 | ···················· | ·A·················· | ····················· | ···················· | ···················· | ···················· |
| F | 8516 | hCoV-19/Switzerland/ZH-ETHZ-190039/2020\|EPI_ISL_50 | ···················· | ···················· | ····················· | ···················· | ···················· | ···················· |
| F | 8517 | hCoV-19/Switzerland/VD-ETHZ-190040/2020\|EPI_ISL_50 | ···················· | ···················· | ····················· | ···················· | ···················· | ···················· |
| F | 8518 | hCoV-19/Switzerland/BE-ETHZ-190041/2020\|EPI_ISL_50 | ···················· | ···················· | ····················· | ···················· | ···················· | ···················· |
| F | 8519 | hCoV-19/Switzerland/UR-ETHZ-190042/2020\|EPI_ISL_50 | ···················· | ···················· | ····················· | ···················· | ···················· | ···················· |
| F | 8520 | hCoV-19/Switzerland/BE-ETHZ-190043/2020\|EPI_ISL_50 | ···················· | ···················· | ····················· | ···················· | ···················· | ···················· |
| F | 8521 | hCoV-19/Switzerland/VD-ETHZ-190044/2020\|EPI_ISL_50 | ···················· | ···················· | ····················· | ···················· | ···················· | ···················· |
| F | 8522 | hCoV-19/Switzerland/BE-ETHZ-190046/2020\|EPI_ISL_50 | ···················· | ···················· | ····················· | ···················· | ···················· | ···················· |
| F | 8523 | hCoV-19/Switzerland/BL-ETHZ-190047/2020\|EPI_ISL_50 | ···················· | ···················· | ····················· | ···················· | ···················· | ···················· |
| F | 8524 | hCoV-19/Switzerland/BL-ETHZ-190049/2020\|EPI_ISL_50 | ···················· | ···················· | ····················· | ···················· | ···················· | ···················· |
| F | 8525 | hCoV-19/Switzerland/BL-ETHZ-190050/2020\|EPI_ISL_50 | ···················· | ···················· | ····················· | ···················· | ···················· | ···················· |
| F | 8526 | hCoV-19/Switzerland/BL-ETHZ-190051/2020\|EPI_ISL_50 | ···················· | ···················· | ····················· | ···················· | ···················· | ···················· |
| F | 8527 | hCoV-19/Switzerland/BL-ETHZ-190052/2020\|EPI_ISL_50 | ···················· | ···················· | ····················· | ···················· | ···················· | ···················· |
| F | 8528 | hCoV-19/Switzerland/ZH-ETHZ-190053/2020\|EPI_ISL_50 | ···················· | ···················· | ····················· | ···················· | ···················· | ···················· |
| F | 8529 | hCoV-19/Switzerland/BL-ETHZ-190055/2020\|EPI_ISL_50 | ···················· | ···················· | ····················· | ···················· | ···················· | ···················· |
| F | 8530 | hCoV-19/Switzerland/ZH-ETHZ-190056/2020\|EPI_ISL_50 | ···················· | ···················· | ····················· | ···················· | ···················· | ···················· |
| F | 8531 | hCoV-19/Switzerland/AG-ETHZ-190058/2020\|EPI_ISL_50 | ···················· | ···················· | ····················· | ···················· | ···················· | ···················· |
| F | 8532 | hCoV-19/Switzerland/BL-ETHZ-190059/2020\|EPI_ISL_50 | ···················· | ···················· | ····················· | ···················· | ···················· | ···················· |
| F | 8533 | hCoV-19/Switzerland/BL-ETHZ-190060/2020\|EPI_ISL_50 | ···················· | ···················· | ····················· | ···················· | ···················· | ···················· |
| F | 8534 | hCoV-19/Switzerland/BL-ETHZ-190061/2020\|EPI_ISL_50 | ···················· | ···················· | ····················· | ···················· | ···················· | ···················· |
| F | 8535 | hCoV-19/Switzerland/BE-ETHZ-190063/2020\|EPI_ISL_50 | ···················· | ·A·················· | ····················· | ···················· | ···················· | ···················· |
| F | 8536 | hCoV-19/Switzerland/LU-ETHZ-190064/2020\|EPI_ISL_50 | ···················· | ···················· | ····················· | ···················· | ···················· | ···················· |
| F | 8537 | hCoV-19/Switzerland/BE-ETHZ-190065/2020\|EPI_ISL_50 | ···················· | ···················· | ····················· | ···················· | ···················· | ···················· |
| F | 8538 | hCoV-19/Switzerland/SZ-ETHZ-190068/2020\|EPI_ISL_50 | ···················· | ·A·················· | ····················· | ···················· | ···················· | ···················· |
| F | 8539 | hCoV-19/Switzerland/BL-ETHZ-190071/2020\|EPI_ISL_50 | ···················· | ···················· | ····················· | ···················· | ···················· | ···················· |
| F | 8540 | hCoV-19/Switzerland/BE-ETHZ-190074/2020\|EPI_ISL_50 | ···················· | ···················· | ····················· | ···················· | ···················· | ···················· |
| F | 8541 | hCoV-19/Switzerland/BL-ETHZ-190075/2020\|EPI_ISL_50 | ···················· | ···················· | ····················· | ···················· | ···················· | ···················· |
| F | 8542 | hCoV-19/Switzerland/BL-ETHZ-190076/2020\|EPI_ISL_50 | ···················· | ···················· | ····················· | ···················· | ···················· | ···················· |
| F | 8543 | hCoV-19/Switzerland/BL-ETHZ-190078/2020\|EPI_ISL_50 | ···················· | ···················· | ····················· | ···················· | ···················· | ···················· |
| F | 8544 | hCoV-19/Switzerland/BE-ETHZ-190079/2020\|EPI_ISL_50 | ···················· | ·A·················· | ····················· | ···················· | ···················· | ···················· |
| F | 8545 | hCoV-19/Switzerland/AG-ETHZ-190081/2020\|EPI_ISL_50 | ···················· | ·A·················· | ····················· | ···················· | ···················· | ···················· |
| F | 8546 | hCoV-19/Switzerland/AG-ETHZ-190082/2020\|EPI_ISL_50 | ···················· | ···················· | ····················· | ···················· | ···················· | ···················· |
| F | 8547 | hCoV-19/Switzerland/BL-ETHZ-190083/2020\|EPI_ISL_50 | ···················· | ···················· | ····················· | ···················· | ···················· | ···················· |
| F | 8548 | hCoV-19/Switzerland/AG-ETHZ-190084/2020\|EPI_ISL_50 | ···················· | ···················· | ····················· | ···················· | ···················· | ···················· |
| F | 8549 | hCoV-19/Switzerland/BS-ETHZ-190085/2020\|EPI_ISL_50 | ···················· | ···················· | ····················· | ···················· | ···················· | ···················· |
| F | 8550 | hCoV-19/USA/OR-OHSU-0310/2020\|EPI_ISL_509101\|2020- | ···················· | ···················· | ····················· | ···················· | ···················· | ···················· |
| F | 8551 | hCoV-19/USA/OR-OHSU-0323/2020\|EPI_ISL_509102\|2020- | ···················· | ···················· | ····················· | ···················· | ···················· | ···················· |
| F | 8552 | hCoV-19/USA/WA-OHSU-0327/2020\|EPI_ISL_509103\|2020- | ···················· | ···················· | ····················· | ···················· | ···················· | ···················· |
| F | 8553 | hCoV-19/USA/OR-OHSU-0332/2020\|EPI_ISL_509104\|2020- | ···················· | ···················· | ····················· | ···················· | ···················· | ···················· |
| F | 8554 | hCoV-19/USA/OR-OHSU-0334/2020\|EPI_ISL_509106\|2020- | ···················· | ···················· | ····················· | ···················· | ···················· | ···················· |
| F | 8555 | hCoV-19/USA/OR-OHSU-0335/2020\|EPI_ISL_509107\|2020- | ···················· | ···················· | ····················· | ···················· | ···················· | ···················· |
| F | 8556 | hCoV-19/USA/OR-OHSU-0336/2020\|EPI_ISL_509108\|2020- | ···················· | ···················· | ····················· | ···················· | ···················· | ···················· |
| F | 8557 | hCoV-19/USA/OR-OHSU-0342/2020\|EPI_ISL_509109\|2020- | ···················· | ···················· | ····················· | ···················· | ···················· | ···················· |
| F | 8558 | hCoV-19/USA/OR-OHSU-0343/2020\|EPI_ISL_509110\|2020- | ···················· | ···················· | ····················· | ···················· | ···················· | ···················· |
| F | 8559 | hCoV-19/USA/OR-OHSU-0344/2020\|EPI_ISL_509111\|2020- | ···················· | ···················· | ····················· | ···················· | ···················· | ···················· |
| F | 8560 | hCoV-19/USA/OR-OHSU-0347/2020\|EPI_ISL_509112\|2020- | ···················· | ···················· | ····················· | ···················· | ···················· | ···················· |
| F | 8561 | hCoV-19/USA/OR-OHSU-0348/2020\|EPI_ISL_509113\|2020- | ···················· | ···················· | ····················· | ···················· | ···················· | ···················· |
| F | 8562 | hCoV-19/USA/OR-OHSU-0349/2020\|EPI_ISL_509114\|2020- | ···················· | ···················· | ····················· | ···················· | ···················· | ···················· |

| | | | | | | | | |
|---|---|---|---|---|---|---|---|---|
| F | 8563 | hCoV-19/USA/OR-OHSU-0353/2020\|EPI_ISL_509116\|2020- | .................... | .................... | ..................... | .................... | .................... | .................... |
| F | 8564 | hCoV-19/USA/OR-OHSU-0360/2020\|EPI_ISL_509117\|2020- | .................... | .................... | ..................... | .................... | .................... | .................... |
| F | 8565 | hCoV-19/USA/WA-OHSU-0366/2020\|EPI_ISL_509118\|2020- | .................... | .................... | ..................... | .................... | .................... | .................... |
| F | 8566 | hCoV-19/USA/OR-OHSU-0367/2020\|EPI_ISL_509119\|2020- | .................... | .................... | ..................... | .................... | .................... | .................... |
| F | 8567 | hCoV-19/USA/OR-OHSU-0369/2020\|EPI_ISL_509120\|2020- | .................... | .................... | ..................... | .................... | .................... | .................... |
| F | 8568 | hCoV-19/USA/OR-OHSU-0380/2020\|EPI_ISL_509121\|2020- | .................... | .................... | ..................... | .................... | .................... | .................... |
| F | 8569 | hCoV-19/USA/OR-OHSU-0382/2020\|EPI_ISL_509122\|2020- | .................... | .................... | ..................... | .................... | .................... | .................... |
| F | 8570 | hCoV-19/USA/OR-OHSU-0385/2020\|EPI_ISL_509123\|2020- | .................... | .................... | ..................... | .................... | .................... | .................... |
| F | 8571 | hCoV-19/USA/WA-OHSU-0386/2020\|EPI_ISL_509124\|2020- | .................... | .................... | ..................... | .................... | .................... | .................... |
| F | 8572 | hCoV-19/USA/OR-OHSU-0387/2020\|EPI_ISL_509125\|2020- | .................... | .................... | ..................... | .................... | .................... | .................... |
| F | 8573 | hCoV-19/USA/OR-OHSU-0389/2020\|EPI_ISL_509126\|2020- | .................... | .................... | ..................... | .................... | .................... | .................... |
| F | 8574 | hCoV-19/USA/OR-OHSU-0392/2020\|EPI_ISL_509127\|2020- | .................... | .................... | ..................... | .................... | .................... | .................... |
| F | 8575 | hCoV-19/USA/OR-OHSU-0393/2020\|EPI_ISL_509128\|2020- | .................... | .................... | ..................... | .................... | .................... | .................... |
| F | 8576 | hCoV-19/USA/OR-OHSU-0396/2020\|EPI_ISL_509129\|2020- | .................... | .................... | ..................... | .................... | .................... | .................... |
| F | 8577 | hCoV-19/USA/OR-OHSU-0398/2020\|EPI_ISL_509130\|2020- | .................... | .................... | ..................... | .................... | .................... | .................... |
| F | 8578 | hCoV-19/USA/OR-OHSU-0415/2020\|EPI_ISL_509131\|2020- | .................... | .................... | ..................... | .................... | .................... | .................... |
| F | 8579 | hCoV-19/USA/OR-OHSU-0418/2020\|EPI_ISL_509132\|2020- | .................... | .................... | ..................... | .................... | .................... | .................... |
| F | 8580 | hCoV-19/USA/OR-OHSU-0419/2020\|EPI_ISL_509133\|2020- | .................... | .A.................. | ..................... | .................... | .................... | .................... |
| F | 8581 | hCoV-19/USA/OR-OHSU-0422/2020\|EPI_ISL_509134\|2020- | .................... | .................... | ..................... | .................... | .................... | .................... |
| F | 8582 | hCoV-19/USA/OR-OHSU-0423/2020\|EPI_ISL_509135\|2020- | .................... | .................... | ..................... | .................... | .................... | .................... |
| F | 8583 | hCoV-19/USA/OR-OHSU-0433/2020\|EPI_ISL_509136\|2020- | .................... | .................... | ..................... | .................... | .................... | .................... |
| F | 8584 | hCoV-19/USA/OR-OHSU-0434/2020\|EPI_ISL_509137\|2020- | .................... | .................... | ..................... | .................... | .................... | .................... |
| F | 8585 | hCoV-19/USA/WA-OHSU-0437/2020\|EPI_ISL_509138\|2020- | .................... | .................... | ..................... | .................... | .................... | .................... |
| F | 8586 | hCoV-19/USA/OR-OHSU-0438/2020\|EPI_ISL_509139\|2020- | .................... | .................... | ..................... | .................... | .................... | .................... |
| F | 8594 | hCoV-19/Portugal/PT1455/2020\|EPI_ISL_511414\|2020-0 | .................... | .................... | ..................... | .................... | .................... | .................... |
| F | 8595 | hCoV-19/Portugal/PT1456/2020\|EPI_ISL_511415\|2020-0 | .................... | .................... | ..................... | .................... | .................... | .................... |
| F | 8596 | hCoV-19/Portugal/PT1457/2020\|EPI_ISL_511416\|2020-0 | .................... | .................... | ..................... | .................... | .................... | .................... |
| F | 8597 | hCoV-19/Portugal/PT1458/2020\|EPI_ISL_511417\|2020-0 | .................... | .................... | ..................... | .................... | .................... | .................... |
| F | 8598 | hCoV-19/Portugal/PT1459/2020\|EPI_ISL_511418\|2020-0 | .................... | .................... | ..................... | .................... | .................... | .................... |
| F | 8599 | hCoV-19/Portugal/PT1460/2020\|EPI_ISL_511419\|2020-0 | .................... | .................... | ..................... | .................... | .................... | .................... |
| F | 8600 | hCoV-19/Portugal/PT1466/2020\|EPI_ISL_511425\|2020-0 | .................... | .................... | ..................... | .................... | .................... | .................... |
| F | 8601 | hCoV-19/Portugal/PT1467/2020\|EPI_ISL_511426\|2020-0 | .................... | .................... | ..................... | .................... | .................... | .................... |
| F | 8602 | hCoV-19/Portugal/PT1468/2020\|EPI_ISL_511427\|2020-0 | .................... | .................... | ..................... | .................... | .................... | .................... |
| F | 8603 | hCoV-19/Portugal/PT1469/2020\|EPI_ISL_511428\|2020-0 | .................... | .................... | ..................... | .................... | .................... | .................... |
| F | 8604 | hCoV-19/Portugal/PT1470/2020\|EPI_ISL_511429\|2020-0 | .................... | .................... | ..................... | .................... | .................... | .................... |
| F | 8605 | hCoV-19/Portugal/PT1471/2020\|EPI_ISL_511430\|2020-0 | .................... | .................... | ..................... | .................... | .................... | .................... |
| F | 8606 | hCoV-19/Portugal/PT1472/2020\|EPI_ISL_511431\|2020-0 | .................... | .................... | ..................... | .................... | .................... | .................... |
| F | 8607 | hCoV-19/Portugal/PT1473/2020\|EPI_ISL_511432\|2020-0 | .................... | .................... | ..................... | .................... | .................... | .................... |
| F | 8608 | hCoV-19/Portugal/PT1474/2020\|EPI_ISL_511433\|2020-0 | .................... | .................... | ..................... | .................... | .................... | .................... |
| F | 8609 | hCoV-19/Portugal/PT1475/2020\|EPI_ISL_511434\|2020-0 | .................... | .................... | ..................... | .................... | .................... | .................... |
| F | 8610 | hCoV-19/Portugal/PT1476/2020\|EPI_ISL_511435\|2020-0 | .................... | .................... | ..................... | .................... | .................... | .................... |
| F | 8611 | hCoV-19/Portugal/PT1477/2020\|EPI_ISL_511436\|2020-0 | .................... | .................... | ..................... | .................... | .................... | .................... |
| F | 8612 | hCoV-19/Portugal/PT1478a/2020\|EPI_ISL_511437\|2020- | .................... | .................... | ..................... | .................... | .................... | .................... |
| F | 8613 | hCoV-19/Portugal/PT1479/2020\|EPI_ISL_511438\|2020-0 | .................... | .................... | ..................... | .................... | .................... | .................... |
| F | 8614 | hCoV-19/Portugal/PT1480/2020\|EPI_ISL_511439\|2020-0 | .................... | .................... | ..................... | .................... | .................... | .................... |
| F | 8615 | hCoV-19/Portugal/PT1482/2020\|EPI_ISL_511441\|2020-0 | .................... | .................... | ..................... | .................... | .................... | .................... |
| F | 8616 | hCoV-19/Portugal/PT1483/2020\|EPI_ISL_511442\|2020-0 | .................... | .................... | ..................... | .................... | .................... | .................... |
| F | 8617 | hCoV-19/Portugal/PT1485/2020\|EPI_ISL_511444\|2020-0 | .................... | .................... | ..................... | .................... | .................... | .................... |
| F | 8618 | hCoV-19/Portugal/PT1487/2020\|EPI_ISL_511446\|2020-0 | .................... | .................... | ..................... | .................... | .................... | .................... |
| F | 8619 | hCoV-19/Portugal/PT1488/2020\|EPI_ISL_511447\|2020-0 | .................... | .................... | ..................... | .................... | .................... | .................... |
| F | 8620 | hCoV-19/Portugal/PT1489/2020\|EPI_ISL_511448\|2020-0 | .................... | .................... | ..................... | .................... | .................... | .................... |
| F | 8621 | hCoV-19/Portugal/PT1490/2020\|EPI_ISL_511449\|2020-0 | .................... | .................... | ..................... | .................... | .................... | .................... |
| F | 8622 | hCoV-19/Portugal/PT1491/2020\|EPI_ISL_511450\|2020-0 | .................... | .................... | ..................... | .................... | .................... | .................... |
| F | 8623 | hCoV-19/Portugal/PT1492/2020\|EPI_ISL_511451\|2020-0 | .................... | .................... | ..................... | .................... | .................... | .................... |
| F | 8624 | hCoV-19/Portugal/PT1493/2020\|EPI_ISL_511452\|2020-0 | .................... | .................... | ..................... | .................... | .................... | .................... |
| F | 8625 | hCoV-19/Portugal/PT1494/2020\|EPI_ISL_511453\|2020-0 | .................... | .................... | ..................... | .................... | .................... | .................... |
| F | 8626 | hCoV-19/Portugal/PT1496/2020\|EPI_ISL_511455\|2020-0 | .................... | .................... | ..................... | .................... | .................... | .................... |
| F | 8627 | hCoV-19/Portugal/PT1497/2020\|EPI_ISL_511456\|2020-0 | .................... | .................... | ..................... | .................... | .................... | .................... |
| F | 8628 | hCoV-19/Portugal/PT1498/2020\|EPI_ISL_511457\|2020-0 | .................... | .................... | ..................... | .................... | .................... | .................... |
| F | 8629 | hCoV-19/Australia/WA94/2020\|EPI_ISL_512747\|2020-04 | .................... | .................... | ..................... | .................... | .................... | .................... |
| F | 8630 | hCoV-19/Australia/WA95/2020\|EPI_ISL_512748\|2020-04 | .................... | .................... | ..................... | .................... | .................... | .................... |
| F | 8631 | hCoV-19/Australia/WA96/2020\|EPI_ISL_512749\|2020-03 | .................... | .................... | ..................... | .................... | .................... | .................... |
| F | 8632 | hCoV-19/Australia/WA103/2020\|EPI_ISL_512756\|2020-0 | .................... | .................... | ..................... | .................... | .................... | .................... |
| F | 8633 | hCoV-19/Australia/WA104/2020\|EPI_ISL_512757\|2020-0 | .................... | .................... | ..................... | .................... | .................... | .................... |
| F | 8634 | hCoV-19/Australia/WA105/2020\|EPI_ISL_512758\|2020-0 | .................... | .................... | ..................... | .................... | .................... | .................... |
| F | 8635 | hCoV-19/Australia/WA106/2020\|EPI_ISL_512759\|2020-0 | .................... | .................... | ..................... | .................... | .................... | .................... |
| F | 8636 | hCoV-19/Australia/WA107/2020\|EPI_ISL_512760\|2020-0 | .................... | .............A...... | ..................... | .................... | .................... | .................... |
| F | 8637 | hCoV-19/Australia/WA108/2020\|EPI_ISL_512761\|2020-0 | .................... | .................... | ..................... | .................... | .................... | .................... |
| F | 8638 | hCoV-19/Australia/WA109/2020\|EPI_ISL_512762\|2020-0 | .................... | .................... | ..................... | .................... | .................... | .................... |
| F | 8639 | hCoV-19/Australia/WA110/2020\|EPI_ISL_512763\|2020-0 | .................... | .................... | ..................... | .................... | .................... | .................... |
| F | 8640 | hCoV-19/Australia/WA111/2020\|EPI_ISL_512764\|2020-0 | .................... | .................... | ..................... | .................... | .................... | .................... |
| F | 8641 | hCoV-19/Australia/WA112/2020\|EPI_ISL_512765\|2020-0 | .................... | .................... | ..................... | .................... | .................... | .................... |
| F | 8642 | hCoV-19/USA/FL-USAFSAM-S009/2020\|EPI_ISL_512785\|20 | .................... | .................... | ..................... | .................... | .................... | .................... |
| F | 8643 | hCoV-19/USA/FL-USAFSAM-S011/2020\|EPI_ISL_512787\|20 | .................... | .................... | ..................... | .................... | .................... | .................... |
| F | 8644 | hCoV-19/SaudiArabia/KAUST-JEDDAH1031/2020\|EPI_ISL_ | .................... | .A.................. | ..................... | .................... | .................... | ..A................. |
| F | 8645 | hCoV-19/SaudiArabia/KAUST-JEDDAH1032/2020\|EPI_ISL_ | .................... | .................... | ..................... | .................... | .................... | .................... |
| F | 8646 | hCoV-19/SaudiArabia/KAUST-JEDDAH1033/2020\|EPI_ISL_ | .................... | .................... | ..................... | .................... | .................... | .................... |
| F | 8647 | hCoV-19/SaudiArabia/KAUST-JEDDAH1034/2020\|EPI_ISL_ | .................... | .................... | ..................... | .................... | .................... | .................... |
| F | 8648 | hCoV-19/SaudiArabia/KAUST-JEDDAH1054/2020\|EPI_ISL_ | .................... | .................... | ..................... | .................... | .................... | .................... |
| F | 8649 | hCoV-19/SaudiArabia/KAUST-JEDDAH1055/2020\|EPI_ISL_ | .................... | .................... | ..................... | .................... | .................... | .................... |

| | | | | | | | | |
|---|---|---|---|---|---|---|---|---|
| F | 8650 | hCoV-19/SaudiArabia/KAUST-JEDDAH1057/2020\|EPI_ISL_ | ···················· | ···················· | ····················· | ···················· | ···················· | ···················· |
| F | 8651 | hCoV-19/SaudiArabia/KAUST-JEDDAH1058/2020\|EPI_ISL_ | ···················· | ···················· | ····················· | ···················· | ···················· | ···················· |
| F | 8652 | hCoV-19/SaudiArabia/KAUST-JEDDAH313/2020\|EPI_ISL_5 | ···················· | ···················· | ····················· | ···················· | ···················· | ···················· |
| F | 8653 | hCoV-19/SaudiArabia/KAUST-JEDDAH317/2020\|EPI_ISL_5 | ···················· | ···················· | ····················· | ···················· | ···················· | ···················· |
| F | 8654 | hCoV-19/SaudiArabia/KAUST-JEDDAH456/2020\|EPI_ISL_5 | ···················· | ···················· | ····················· | ···················· | ···················· | ···················· |
| F | 8655 | hCoV-19/SaudiArabia/KAUST-JEDDAH457/2020\|EPI_ISL_5 | ···················· | ···················· | ····················· | ···················· | ···················· | ···················· |
| F | 8656 | hCoV-19/SaudiArabia/KAUST-JEDDAH462/2020\|EPI_ISL_5 | ···················· | ···················· | ····················· | ···················· | ···················· | ···················· |
| F | 8657 | hCoV-19/South Africa/LR00027/2020\|EPI_ISL_515156\|2 | ···················· | ···················· | ····················· | ···················· | ···················· | ···················· |
| F | 8658 | hCoV-19/South Africa/LR00039/2020\|EPI_ISL_515166\|2 | ···················· | ···················· | ····················· | ···················· | ···················· | ···················· |
| F | 8659 | hCoV-19/South Africa/LR00045/2020\|EPI_ISL_515172\|2 | ···················· | ···················· | ····················· | ···················· | ···················· | ···················· |
| F | 8660 | hCoV-19/Ghana/KATH23/2020\|EPI_ISL_515181\|2020-03-2 | ···················· | ···················· | ····················· | ··M················· | ···················· | ···················· |
| F | 8661 | hCoV-19/Ghana/TTH10/2020\|EPI_ISL_515182\|2020-03-27 | ···················· | ···················· | ····················· | ···················· | ···················· | ···················· |
| F | 8662 | hCoV-19/Ghana/TTH11/2020\|EPI_ISL_515183\|2020-03-27 | ···················· | ···················· | ····················· | ···················· | ···················· | ···················· |
| F | 8663 | hCoV-19/Latvia/098/2020\|EPI_ISL_515185\|2020-07-29 | ···················· | ···················· | ····················· | ···················· | ···················· | ···················· |
| F | 8664 | hCoV-19/Latvia/099/2020\|EPI_ISL_515186\|2020-07-31 | ···················· | ···················· | ····················· | ···················· | ···················· | ···················· |
| F | 8665 | hCoV-19/Latvia/100/2020\|EPI_ISL_515187\|2020-08-04 | ···················· | ···················· | ····················· | ···················· | ···················· | ···················· |
| F | 8666 | hCoV-19/Latvia/101/2020\|EPI_ISL_515188\|2020-08-04 | ···················· | ···················· | ····················· | ···················· | ···················· | ···················· |
| F | 8667 | hCoV-19/Latvia/102/2020\|EPI_ISL_515189\|2020-08-05 | ···················· | ···················· | ····················· | ···················· | ···················· | ···················· |
| F | 8668 | hCoV-19/Latvia/103/2020\|EPI_ISL_515190\|2020-08-05 | ···················· | ···················· | ····················· | ···················· | ···················· | ···················· |
| F | 8669 | hCoV-19/Latvia/105/2020\|EPI_ISL_515192\|2020-08-05 | ···················· | ···················· | ····················· | ···················· | ···················· | ···················· |
| F | 8670 | hCoV-19/Latvia/106/2020\|EPI_ISL_515193\|2020-08-05 | ···················· | ···················· | ····················· | ···················· | ···················· | ···················· |
| F | 8671 | hCoV-19/Latvia/107/2020\|EPI_ISL_515194\|2020-08-05 | ···················· | ···················· | ····················· | ···················· | ···················· | ···················· |
| F | 8672 | hCoV-19/Latvia/108/2020\|EPI_ISL_515195\|2020-08-05 | ···················· | ···················· | ····················· | ···················· | ···················· | ···················· |
| F | 8673 | hCoV-19/Latvia/109/2020\|EPI_ISL_515196\|2020-08-03 | ···················· | ···················· | ····················· | ···················· | ···················· | ···················· |
| F | 8674 | hCoV-19/Canada/QC_AA1/2020\|EPI_ISL_515197\|2020-04- | ···················· | ···················· | ····················· | ···················· | ···················· | ···················· |
| F | 8675 | hCoV-19/USA/WI-UW-1044/2020\|EPI_ISL_516470\|2020-08 | ···················· | ···················· | ····················· | ···················· | ···················· | ···················· |
| F | 8676 | hCoV-19/USA/WI-UW-1045/2020\|EPI_ISL_516471\|2020-07 | ···················· | ···················· | ····················· | ···················· | ···················· | ···················· |
| F | 8677 | hCoV-19/USA/WI-UW-1046/2020\|EPI_ISL_516472\|2020-07 | ···················· | ···················· | ····················· | ···················· | ···················· | ···················· |
| F | 8678 | hCoV-19/USA/WI-UW-1047/2020\|EPI_ISL_516473\|2020-08 | ···················· | ···················· | ····················· | ···················· | ···················· | ···················· |
| F | 8679 | hCoV-19/USA/WI-UW-1048/2020\|EPI_ISL_516474\|2020-07 | ···················· | ···················· | ····················· | ···················· | ···················· | ···················· |
| F | 8680 | hCoV-19/USA/WI-UW-1049/2020\|EPI_ISL_516475\|2020-08 | ···················· | ·············A······ | ····················· | ···················· | ···················· | ···················· |
| F | 8681 | hCoV-19/USA/WI-UW-1050/2020\|EPI_ISL_516476\|2020-07 | ···················· | ···················· | ····················· | ···················· | ···················· | ···················· |
| F | 8682 | hCoV-19/USA/WI-UW-1051/2020\|EPI_ISL_516477\|2020-08 | ···················· | ···················· | ····················· | ···················· | ···················· | ···················· |
| F | 8683 | hCoV-19/USA/WI-UW-1052/2020\|EPI_ISL_516478\|2020-08 | ···················· | ···················· | ····················· | ···················· | ···················· | ···················· |
| F | 8684 | hCoV-19/USA/WI-UW-1053/2020\|EPI_ISL_516479\|2020-07 | ···················· | ···················· | ····················· | ···················· | ···················· | ···················· |
| F | 8685 | hCoV-19/USA/WI-UW-1054/2020\|EPI_ISL_516480\|2020-07 | ···················· | ···················· | ····················· | ···················· | ···················· | ···················· |
| F | 8686 | hCoV-19/USA/WI-UW-1055/2020\|EPI_ISL_516481\|2020-07 | ···················· | ···················· | ····················· | ···················· | ···················· | ···················· |
| F | 8687 | hCoV-19/USA/WI-UW-1056/2020\|EPI_ISL_516482\|2020-07 | ···················· | ···················· | ····················· | ···················· | ···················· | ···················· |
| F | 8688 | hCoV-19/USA/WI-UW-1057/2020\|EPI_ISL_516483\|2020-07 | ···················· | ···················· | ····················· | ···················· | ···················· | ···················· |
| F | 8689 | hCoV-19/USA/WI-UW-1058/2020\|EPI_ISL_516484\|2020-07 | ···················· | ···················· | ····················· | ···················· | ···················· | ···················· |
| F | 8690 | hCoV-19/USA/WI-UW-1059/2020\|EPI_ISL_516485\|2020-07 | ···················· | ···················· | ····················· | ···················· | ···················· | ···················· |
| F | 8691 | hCoV-19/USA/WI-UW-1060/2020\|EPI_ISL_516486\|2020-07 | ···················· | ···················· | ····················· | ···················· | ···················· | ···················· |
| F | 8692 | hCoV-19/USA/WI-UW-1061/2020\|EPI_ISL_516487\|2020-07 | ···················· | ·A·················· | ····················· | ···················· | ···················· | ···················· |
| F | 8693 | hCoV-19/USA/WI-UW-1062/2020\|EPI_ISL_516488\|2020-07 | ···················· | ···················· | ····················· | ···················· | ···················· | ···················· |
| F | 8694 | hCoV-19/USA/WI-UW-1063/2020\|EPI_ISL_516489\|2020-07 | ···················· | ···················· | ····················· | ···················· | ···················· | ···················· |
| F | 8695 | hCoV-19/USA/IL-UW-1064/2020\|EPI_ISL_516490\|2020-07 | ···················· | ···················· | ····················· | ···················· | ···················· | ···················· |
| F | 8696 | hCoV-19/USA/IL-UW-1065/2020\|EPI_ISL_516491\|2020-04 | ···················· | ···················· | ····················· | ···················· | ···················· | ···················· |
| F | 8697 | hCoV-19/USA/WI-UW-1066/2020\|EPI_ISL_516492\|2020-07 | ···················· | ·············A······ | ····················· | ···················· | ···················· | ···················· |
| F | 8698 | hCoV-19/USA/WI-UW-1067/2020\|EPI_ISL_516493\|2020-07 | ···················· | ···················· | ····················· | ···················· | ···················· | ···················· |
| F | 8699 | hCoV-19/USA/WI-UW-1068/2020\|EPI_ISL_516494\|2020-07 | ···················· | ···················· | ····················· | ···················· | ···················· | ···················· |
| F | 8700 | hCoV-19/USA/WI-UW-1069/2020\|EPI_ISL_516495\|2020-07 | ···················· | ·············A······ | ····················· | ···················· | ···················· | ···················· |
| F | 8701 | hCoV-19/USA/WI-UW-1070/2020\|EPI_ISL_516496\|2020-08 | ···················· | ·············A······ | ····················· | ···················· | ···················· | ···················· |
| F | 8702 | hCoV-19/USA/WI-UW-1071/2020\|EPI_ISL_516497\|2020-07 | ···················· | ···················· | ····················· | ···················· | ···················· | ···················· |
| F | 8703 | hCoV-19/USA/WI-UW-1072/2020\|EPI_ISL_516498\|2020-07 | ···················· | ···················· | ····················· | ···················· | ···················· | ···················· |
| F | 8704 | hCoV-19/USA/WI-UW-1073/2020\|EPI_ISL_516499\|2020-07 | ···················· | ···················· | ····················· | ···················· | ···················· | ···················· |
| F | 8705 | hCoV-19/USA/WA-S2319/2020\|EPI_ISL_496951\|2020-06-1 | ···················· | ···················· | ····················· | ···················· | ···················· | ···················· |
| F | 8706 | hCoV-19/USA/WA-S2320/2020\|EPI_ISL_496952\|2020-06-1 | ···················· | ···················· | ····················· | ···················· | ···················· | ···················· |
| F | 8707 | hCoV-19/USA/WA-S2324/2020\|EPI_ISL_496956\|2020-06-1 | ···················· | ···················· | ····················· | ···················· | ···················· | ···················· |
| F | 8708 | hCoV-19/USA/WA-S2328/2020\|EPI_ISL_496960\|2020-06-1 | ···················· | ···················· | ····················· | ···················· | ···················· | ···················· |
| F | 8709 | hCoV-19/USA/WA-S2329/2020\|EPI_ISL_496961\|2020-06-1 | ···················· | ···················· | ····················· | ···················· | ···················· | ···················· |
| F | 8710 | hCoV-19/USA/WA-S2331/2020\|EPI_ISL_496963\|2020-06-1 | ···················· | ···················· | ····················· | ···················· | ···················· | ···················· |
| F | 8711 | hCoV-19/USA/WA-S2332/2020\|EPI_ISL_496964\|2020-06-1 | ···················· | ···················· | ····················· | ···················· | ···················· | ···················· |
| F | 8712 | hCoV-19/USA/WA-S2333/2020\|EPI_ISL_496965\|2020-06-1 | ···················· | ···················· | ····················· | ···················· | ···················· | ···················· |
| F | 8713 | hCoV-19/USA/WA-S2334/2020\|EPI_ISL_496966\|2020-06-1 | ···················· | ···················· | ····················· | ···················· | ···················· | ···················· |
| F | 8714 | hCoV-19/USA/WA-S2335/2020\|EPI_ISL_496967\|2020-06-1 | ···················· | ···················· | ····················· | ···················· | ···················· | ···················· |
| F | 8715 | hCoV-19/USA/WA-S2336/2020\|EPI_ISL_496968\|2020-06-1 | ···················· | ···················· | ····················· | ···················· | ···················· | ···················· |
| F | 8716 | hCoV-19/USA/WA-S2337/2020\|EPI_ISL_496969\|2020-06-1 | ···················· | ···················· | ····················· | ···················· | ···················· | ···················· |
| F | 8717 | hCoV-19/USA/WA-S2338/2020\|EPI_ISL_496970\|2020-06-1 | ···················· | ···················· | ····················· | ···················· | ···················· | ···················· |
| F | 8718 | hCoV-19/USA/WA-S2361/2020\|EPI_ISL_496993\|2020-06-1 | ···················· | ···················· | ····················· | ···················· | ···················· | ···················· |
| F | 8719 | hCoV-19/USA/WA-S2362/2020\|EPI_ISL_496994\|2020-06-1 | ···················· | ···················· | ····················· | ···················· | ···················· | ···················· |
| F | 8720 | hCoV-19/USA/WA-S2363/2020\|EPI_ISL_496995\|2020-06-1 | ···················· | ···················· | ····················· | ···················· | ···················· | ···················· |
| F | 8721 | hCoV-19/USA/WA-S2364/2020\|EPI_ISL_496996\|2020-06-1 | ···················· | ···················· | ····················· | ···················· | ···················· | ···················· |
| F | 8722 | hCoV-19/USA/WA-S2365/2020\|EPI_ISL_496997\|2020-06-1 | ···················· | ···················· | ····················· | ···················· | ···················· | ···················· |
| F | 8723 | hCoV-19/USA/WA-S2366/2020\|EPI_ISL_496998\|2020-06-1 | ···················· | ···················· | ····················· | ···················· | ···················· | ···················· |
| F | 8724 | hCoV-19/USA/WA-S2367/2020\|EPI_ISL_496999\|2020-06-1 | ···················· | ···················· | ····················· | ···················· | ···················· | ···················· |
| F | 8725 | hCoV-19/Norway/2854/2020\|EPI_ISL_500777\|2020-06-29 | ···················· | ···················· | ····················· | ···················· | ···················· | ···················· |
| F | 8726 | hCoV-19/Norway/2855/2020\|EPI_ISL_500778\|2020-06-25 | ···················· | ···················· | ····················· | ···················· | ···················· | ···················· |
| F | 8727 | hCoV-19/Norway/2856/2020\|EPI_ISL_500779\|2020-07-02 | ···················· | ···················· | ····················· | ···················· | ···················· | ···················· |
| F | 8728 | hCoV-19/South Korea/KCDC2153/2020\|EPI_ISL_506947\|2 | ···················· | ···················· | ····················· | ···················· | ···················· | ···················· |
| F | 8729 | hCoV-19/South Korea/KCDC2176/2020\|EPI_ISL_506970\|2 | ···················· | ···················· | ····················· | ···················· | ···················· | ···················· |

| | | | | | | | | |
|---|---|---|---|---|---|---|---|---|
| F | 8730 | hCoV-19/South Korea/KCDC2177/2020\|EPI_ISL_506971\|2 | ···················· | ···················· | ···················· | ···················· | ···················· | ···················· |
| F | 8731 | hCoV-19/South Korea/KCDC2178/2020\|EPI_ISL_506972\|2 | ···················· | ···················· | ···················· | ···················· | ···················· | ···················· |
| F | 8732 | hCoV-19/South Korea/KCDC2179/2020\|EPI_ISL_506973\|2 | ···················· | ···················· | ···················· | ···················· | ···················· | ···················· |
| F | 8733 | hCoV-19/South Korea/KCDC2180/2020\|EPI_ISL_506974\|2 | ···················· | ···················· | ···················· | ···················· | ···················· | ···················· |
| F | 8734 | hCoV-19/South Korea/KCDC2181/2020\|EPI_ISL_506975\|2 | ···················· | ···················· | ···················· | ···················· | ···················· | ···················· |
| F | 8735 | hCoV-19/South Korea/KCDC2182/2020\|EPI_ISL_506976\|2 | ···················· | ···················· | ···················· | ···················· | ···················· | ···················· |
| F | 8736 | hCoV-19/South Korea/KCDC2183/2020\|EPI_ISL_506977\|2 | ···················· | ···················· | ···················· | ···················· | ···················· | ···················· |
| F | 8737 | hCoV-19/South Korea/KCDC2184/2020\|EPI_ISL_506978\|2 | ···················· | ···················· | ···················· | ···················· | ···················· | ···················· |
| F | 8738 | hCoV-19/South Korea/KCDC2185/2020\|EPI_ISL_506979\|2 | ···················· | ···················· | ···················· | ···················· | ···················· | ···················· |
| F | 8739 | hCoV-19/South Korea/KCDC2190/2020\|EPI_ISL_506984\|2 | ···················· | ···················· | ···················· | ···················· | ···················· | ···················· |
| F | 8740 | hCoV-19/South Korea/KCDC2191/2020\|EPI_ISL_506985\|2 | ···················· | ···················· | ···················· | ···················· | ···················· | ···················· |
| F | 8741 | hCoV-19/South Korea/KCDC2192/2020\|EPI_ISL_506986\|2 | ···················· | ···················· | ···················· | ···················· | ···················· | ···················· |
| F | 8742 | hCoV-19/South Korea/KCDC2193/2020\|EPI_ISL_506987\|2 | ···················· | ···················· | ···················· | ···················· | ···················· | ···················· |
| F | 8743 | hCoV-19/South Korea/KCDC2194/2020\|EPI_ISL_506988\|2 | ···················· | ···················· | ···················· | ···················· | ···················· | ···················· |
| F | 8744 | hCoV-19/South Korea/KCDC2195/2020\|EPI_ISL_506989\|2 | ···················· | ···················· | ···················· | ···················· | ···················· | ···················· |
| F | 8745 | hCoV-19/South Korea/KCDC2196/2020\|EPI_ISL_506990\|2 | ···················· | ···················· | ···················· | ···················· | ···················· | ···················· |
| F | 8746 | hCoV-19/South Korea/KCDC2197/2020\|EPI_ISL_506991\|2 | ···················· | ···················· | ···················· | ···················· | ···················· | ···················· |
| F | 8747 | hCoV-19/South Korea/KCDC2198/2020\|EPI_ISL_506992\|2 | ···················· | ···················· | ···················· | ···················· | ···················· | ···················· |
| F | 8748 | hCoV-19/South Korea/KCDC2199/2020\|EPI_ISL_506993\|2 | ···················· | ···················· | ···················· | ···················· | ···················· | ···················· |
| F | 8749 | hCoV-19/South Korea/KCDC2200/2020\|EPI_ISL_506994\|2 | ···················· | ···················· | ···················· | ···················· | ···················· | ···················· |
| F | 8750 | hCoV-19/South Korea/KCDC2202/2020\|EPI_ISL_506995\|2 | ···················· | ···················· | ···················· | ···················· | ···················· | ···················· |
| F | 8751 | hCoV-19/Malaysia/7685/2020\|EPI_ISL_506996\|2020-04- | ···················· | ···················· | ···················· | ···················· | ···················· | ···················· |
| F | 8752 | hCoV-19/USA/OR-OHSU-0352/2020\|EPI_ISL_509115\|2020- | ···················· | ···················· | ···················· | ···················· | ···················· | ···················· |
| F | 8753 | hCoV-19/USA/OR-OHSU-0439/2020\|EPI_ISL_509140\|2020- | ···················· | ···················· | ···················· | ···················· | ···················· | ···················· |
| F | 8754 | hCoV-19/USA/OR-OHSU-1111/2020\|EPI_ISL_509141\|2020- | ···················· | ···················· | ···················· | ···················· | ···················· | ···················· |
| F | 8755 | hCoV-19/USA/OR-OHSU-1115/2020\|EPI_ISL_509142\|2020- | ···················· | ·A·················· | ···················· | ···················· | ···················· | ···················· |
| F | 8756 | hCoV-19/USA/OR-OHSU-1117/2020\|EPI_ISL_509143\|2020- | ···················· | ···················· | ···················· | ···················· | ···················· | ···················· |
| F | 8757 | hCoV-19/USA/OR-OHSU-1121/2020\|EPI_ISL_509144\|2020- | ···················· | ···················· | ···················· | ···················· | ···················· | ···················· |
| F | 8758 | hCoV-19/USA/OR-OHSU-1127/2020\|EPI_ISL_509145\|2020- | ···················· | ···················· | ···················· | ···················· | ···················· | ···················· |
| F | 8759 | hCoV-19/USA/OR-OHSU-1131/2020\|EPI_ISL_509146\|2020- | ···················· | ···················· | ···················· | ···················· | ···················· | ···················· |
| F | 8760 | hCoV-19/USA/OR-OHSU-1133/2020\|EPI_ISL_509147\|2020- | ···················· | ···················· | ···················· | ···················· | ···················· | ···················· |
| F | 8761 | hCoV-19/USA/OR-OHSU-1135/2020\|EPI_ISL_509148\|2020- | ···················· | ···················· | ···················· | ···················· | ···················· | ···················· |
| F | 8762 | hCoV-19/USA/OR-OHSU-1137/2020\|EPI_ISL_509149\|2020- | ···················· | ···················· | ···················· | ···················· | ···················· | ···················· |
| F | 8763 | hCoV-19/USA/OR-OHSU-1153/2020\|EPI_ISL_509150\|2020- | ···················· | ·A·················· | ···················· | ···················· | ···················· | ···················· |
| F | 8764 | hCoV-19/USA/OR-OHSU-1157/2020\|EPI_ISL_509151\|2020- | ···················· | ···················· | ···················· | ···················· | ···················· | ···················· |
| F | 8765 | hCoV-19/USA/OR-OHSU-1167/2020\|EPI_ISL_509153\|2020- | ···················· | ···················· | ···················· | ···················· | ···················· | ···················· |
| F | 8766 | hCoV-19/USA/OR-OHSU-1169/2020\|EPI_ISL_509154\|2020- | ···················· | ·A·················· | ···················· | ···················· | ···················· | ···················· |
| F | 8767 | hCoV-19/USA/OR-OHSU-1171/2020\|EPI_ISL_509155\|2020- | ···················· | ·A·················· | ···················· | ···················· | ···················· | ···················· |
| F | 8768 | hCoV-19/USA/OR-OHSU-1175/2020\|EPI_ISL_509156\|2020- | ···················· | ···················· | ···················· | ···················· | ···················· | ···················· |
| F | 8769 | hCoV-19/USA/OR-OHSU-1181/2020\|EPI_ISL_509157\|2020- | ···················· | ···················· | ···················· | ···················· | ···················· | ···················· |
| F | 8770 | hCoV-19/USA/OR-OHSU-1185/2020\|EPI_ISL_509159\|2020- | ···················· | ···················· | ···················· | ···················· | ···················· | ···················· |
| F | 8771 | hCoV-19/USA/OR-OHSU-1197/2020\|EPI_ISL_509160\|2020- | ···················· | ···················· | ···················· | ···················· | ···················· | ···················· |
| F | 8772 | hCoV-19/USA/OR-OHSU-1203/2020\|EPI_ISL_509161\|2020- | ···················· | ···················· | ···················· | ···················· | ···················· | ···················· |
| F | 8773 | hCoV-19/USA/OR-OHSU-1205/2020\|EPI_ISL_509162\|2020- | ···················· | ·A·················· | ···················· | ···················· | ···················· | ···················· |
| F | 8774 | hCoV-19/USA/OR-OHSU-1213/2020\|EPI_ISL_509163\|2020- | ···················· | ·A·················· | ···················· | ···················· | ···················· | ···················· |
| F | 8775 | hCoV-19/USA/OR-OHSU-1221/2020\|EPI_ISL_509164\|2020- | ···················· | ·A·················· | ···················· | ···················· | ···················· | ···················· |
| F | 8776 | hCoV-19/USA/OR-OHSU-1223/2020\|EPI_ISL_509165\|2020- | ···················· | ·A·················· | ···················· | ···················· | ···················· | ···················· |
| F | 8777 | hCoV-19/USA/OR-OHSU-1233/2020\|EPI_ISL_509166\|2020- | ···················· | ·A·················· | ···················· | ···················· | ···················· | ···················· |
| F | 8778 | hCoV-19/USA/OR-OHSU-1235/2020\|EPI_ISL_509167\|2020- | ···················· | ·A·················· | ···················· | ···················· | ···················· | ···················· |
| F | 8779 | hCoV-19/USA/OR-OHSU-1239/2020\|EPI_ISL_509168\|2020- | ···················· | ···················· | ···················· | ···················· | ···················· | ···················· |
| F | 8780 | hCoV-19/USA/OR-OHSU-1241/2020\|EPI_ISL_509169\|2020- | ···················· | ···················· | ···················· | ···················· | ···················· | ···················· |
| F | 8781 | hCoV-19/USA/OR-OHSU-1243/2020\|EPI_ISL_509170\|2020- | ···················· | ···················· | ···················· | ···················· | ···················· | ···················· |
| F | 8782 | hCoV-19/USA/OR-OHSU-1245/2020\|EPI_ISL_509171\|2020- | ···················· | ···················· | ···················· | ···················· | ···················· | ···················· |
| F | 8783 | hCoV-19/USA/OR-OHSU-1267/2020\|EPI_ISL_509174\|2020- | ···················· | ···················· | ···················· | ···················· | ···················· | ···················· |
| F | 8784 | hCoV-19/USA/OR-OHSU-1271/2020\|EPI_ISL_509175\|2020- | ···················· | ···················· | ···················· | ···················· | ···················· | ···················· |
| F | 8785 | hCoV-19/USA/OR-OHSU-1273/2020\|EPI_ISL_509176\|2020- | ···················· | ···················· | ···················· | ···················· | ···················· | ···················· |
| F | 8786 | hCoV-19/USA/OR-OHSU-1315/2020\|EPI_ISL_509181\|2020- | ···················· | ·A·················· | ···················· | ···················· | ···················· | ···················· |
| F | 8787 | hCoV-19/USA/OR-OHSU-1317/2020\|EPI_ISL_509182\|2020- | ···················· | ·A·················· | ···················· | ···················· | ···················· | ···················· |
| F | 8788 | hCoV-19/USA/OR-OHSU-1323/2020\|EPI_ISL_509183\|2020- | ···················· | ···················· | ···················· | ···················· | ···················· | ···················· |
| F | 8789 | hCoV-19/USA/OR-OHSU-1331/2020\|EPI_ISL_509184\|2020- | ···················· | ···················· | ···················· | ···················· | ···················· | ···················· |
| F | 8790 | hCoV-19/USA/WA-OHSU-1333/2020\|EPI_ISL_509185\|2020- | ···················· | ···················· | ···················· | ···················· | ···················· | ···················· |
| F | 8791 | hCoV-19/USA/WA-OHSU-1345/2020\|EPI_ISL_509186\|2020- | ···················· | ···················· | ···················· | ···················· | ···················· | ···················· |
| F | 8792 | hCoV-19/USA/OR-OHSU-1349/2020\|EPI_ISL_509187\|2020- | ···················· | ···················· | ···················· | ···················· | ···················· | ···················· |
| F | 8793 | hCoV-19/USA/OR-OHSU-1359/2020\|EPI_ISL_509188\|2020- | ···················· | ···················· | ···················· | ···················· | ···················· | ···················· |
| F | 8794 | hCoV-19/USA/OR-OHSU-1365/2020\|EPI_ISL_509189\|2020- | ···················· | ···················· | ···················· | ···················· | ···················· | ···················· |
| F | 8795 | hCoV-19/USA/OR-OHSU-1389/2020\|EPI_ISL_509191\|2020- | ···················· | ·············A······ | ···················· | ···················· | ···················· | ···················· |
| F | 8796 | hCoV-19/USA/OR-OHSU-1397/2020\|EPI_ISL_509192\|2020- | ···················· | ···················· | ···················· | ···················· | ···················· | ···················· |
| F | 8797 | hCoV-19/USA/OR-OHSU-1401/2020\|EPI_ISL_509193\|2020- | ···················· | ·A·················· | ···················· | ···················· | ···················· | ···················· |
| F | 8798 | hCoV-19/USA/OR-OHSU-1411/2020\|EPI_ISL_509194\|2020- | ···················· | ·A·················· | ···················· | ···················· | ···················· | ···················· |
| F | 8799 | hCoV-19/USA/OR-OHSU-1413/2020\|EPI_ISL_509195\|2020- | ···················· | ·A·················· | ···················· | ···················· | ···················· | ···················· |
| F | 8800 | hCoV-19/USA/OR-OHSU-1425/2020\|EPI_ISL_509197\|2020- | ···················· | ···················· | ···················· | ···················· | ···················· | ···················· |
| F | 8801 | hCoV-19/USA/OR-OHSU-1427/2020\|EPI_ISL_509198\|2020- | ···················· | ···················· | ···················· | ···················· | ···················· | ···················· |
| F | 8802 | hCoV-19/USA/OR-OHSU-1431/2020\|EPI_ISL_509199\|2020- | ···················· | ···················· | ···················· | ···················· | ···················· | ···················· |
| F | 8816 | hCoV-19/Spain/MD-IBV-005283/2020\|EPI_ISL_510169\|20 | ···················· | ···················· | ···················· | ···················· | ···················· | ···················· |
| F | 8837 | hCoV-19/Portugal/PT1441/2020\|EPI_ISL_511400\|2020-0 | ···················· | ···················· | ···················· | ···················· | ···················· | ···················· |
| F | 8838 | hCoV-19/Portugal/PT1442/2020\|EPI_ISL_511401\|2020-0 | ···················· | ···················· | ···················· | ···················· | ···················· | ···················· |
| F | 8839 | hCoV-19/Portugal/PT1443/2020\|EPI_ISL_511402\|2020-0 | ···················· | ···················· | ···················· | ···················· | ···················· | ···················· |
| F | 8840 | hCoV-19/Portugal/PT1444/2020\|EPI_ISL_511403\|2020-0 | ···················· | ···················· | ···················· | ···················· | ···················· | ···················· |
| F | 8841 | hCoV-19/Portugal/PT1445/2020\|EPI_ISL_511404\|2020-0 | ···················· | ···················· | ···················· | ···················· | ···················· | ···················· |
| F | 8842 | hCoV-19/Portugal/PT1446/2020\|EPI_ISL_511405\|2020-0 | ···················· | ···················· | ···················· | ···················· | ···················· | ···················· |

| | | | | | | | |
|---|---|---|---|---|---|---|---|
| F | 8843 hCoV-19/Portugal/PT1447/2020\|EPI_ISL_511406\|2020-0 | ···················· | ···················· | ···················· | ···················· | ···················· | ···················· |
| F | 8844 hCoV-19/Portugal/PT1448/2020\|EPI_ISL_511407\|2020-0 | ···················· | ···················· | ···················· | ···················· | ···················· | ···················· |
| F | 8845 hCoV-19/Portugal/PT1449/2020\|EPI_ISL_511408\|2020-0 | ···················· | ···················· | ···················· | ···················· | ···················· | ···················· |
| F | 8846 hCoV-19/Portugal/PT1450/2020\|EPI_ISL_511409\|2020-0 | ···················· | ···················· | ···················· | ···················· | ···················· | ···················· |
| F | 8847 hCoV-19/Portugal/PT1451/2020\|EPI_ISL_511410\|2020-0 | ···················· | ···················· | ···················· | ···················· | ···················· | ···················· |
| F | 8848 hCoV-19/Portugal/PT1452/2020\|EPI_ISL_511411\|2020-0 | ···················· | ···················· | ···················· | ···················· | ···················· | ···················· |
| F | 8849 hCoV-19/Portugal/PT1453/2020\|EPI_ISL_511412\|2020-0 | ···················· | ···················· | ···················· | ···················· | ···················· | ···················· |
| F | 8850 hCoV-19/Portugal/PT1454/2020\|EPI_ISL_511413\|2020-0 | ···················· | ···················· | ···················· | ···················· | ···················· | ···················· |
| F | 8851 hCoV-19/Portugal/PT1461/2020\|EPI_ISL_511420\|2020-0 | ···················· | ···················· | ···················· | ···················· | ···················· | ···················· |
| F | 8852 hCoV-19/Portugal/PT1463/2020\|EPI_ISL_511422\|2020-0 | ···················· | ···················· | ···················· | ···················· | ···················· | ···················· |
| F | 8853 hCoV-19/Portugal/PT1464/2020\|EPI_ISL_511423\|2020-0 | ···················· | ···················· | ···················· | ···················· | ···················· | ···················· |
| F | 8854 hCoV-19/Portugal/PT1465/2020\|EPI_ISL_511424\|2020-0 | ···················· | ···················· | ···················· | ···················· | ···················· | ···················· |
| F | 8855 hCoV-19/Portugal/PT1499/2020\|EPI_ISL_511458\|2020-0 | ···················· | ···················· | ···················· | ···················· | ···················· | ···················· |
| F | 8856 hCoV-19/Portugal/PT1500/2020\|EPI_ISL_511459\|2020-0 | ···················· | ···················· | ···················· | ···················· | ···················· | ···················· |
| F | 8857 hCoV-19/Portugal/PT1501/2020\|EPI_ISL_511460\|2020-0 | ···················· | ···················· | ···················· | ···················· | ···················· | ···················· |
| F | 8858 hCoV-19/Portugal/PT1502/2020\|EPI_ISL_511461\|2020-0 | ···················· | ···················· | ···················· | ···················· | ···················· | ···················· |
| F | 8859 hCoV-19/Portugal/PT1503/2020\|EPI_ISL_511462\|2020-0 | ···················· | ···················· | ···················· | ···················· | ···················· | ···················· |
| F | 8860 hCoV-19/Portugal/PT1504/2020\|EPI_ISL_511463\|2020-0 | ···················· | ···················· | ···················· | ···················· | ···················· | ···················· |
| F | 8861 hCoV-19/Portugal/PT1506/2020\|EPI_ISL_511465\|2020-0 | ···················· | ···················· | ···················· | ···················· | ···················· | ···················· |
| F | 8862 hCoV-19/Portugal/PT1507/2020\|EPI_ISL_511466\|2020-0 | ···················· | ···················· | ···················· | ···················· | ···················· | ···················· |
| F | 8863 hCoV-19/Portugal/PT1508/2020\|EPI_ISL_511467\|2020-0 | ···················· | ···················· | ···················· | ···················· | ···················· | ···················· |
| F | 8864 hCoV-19/Portugal/PT1510/2020\|EPI_ISL_511469\|2020-0 | ···················· | ···················· | ···················· | ···················· | ···················· | ···················· |
| F | 8865 hCoV-19/Portugal/PT1511/2020\|EPI_ISL_511470\|2020-0 | ···················· | ···················· | ···················· | ···················· | ···················· | ···················· |
| F | 8866 hCoV-19/Portugal/PT1512/2020\|EPI_ISL_511471\|2020-0 | ···················· | ···················· | ···················· | ···················· | ···················· | ···················· |
| F | 8867 hCoV-19/Portugal/PT1513/2020\|EPI_ISL_511472\|2020-0 | ···················· | ···················· | ···················· | ···················· | ···················· | ···················· |
| F | 8868 hCoV-19/Portugal/PT1514/2020\|EPI_ISL_511473\|2020-0 | ···················· | ···················· | ···················· | ···················· | ···················· | ···················· |
| F | 8869 hCoV-19/Portugal/PT1515/2020\|EPI_ISL_511474\|2020-0 | ···················· | ···················· | ···················· | ···················· | ···················· | ···················· |
| F | 8870 hCoV-19/Portugal/PT1518/2020\|EPI_ISL_511477\|2020-0 | ···················· | ···················· | ···················· | ···················· | ···················· | ···················· |
| F | 8871 hCoV-19/Portugal/PT1520/2020\|EPI_ISL_511479\|2020-0 | ···················· | ···················· | ···················· | ···················· | ···················· | ···················· |
| F | 8872 hCoV-19/Portugal/PT0568a/2020\|EPI_ISL_511481\|2020- | ···················· | ···················· | ···················· | ···················· | ···················· | ···················· |
| F | 8873 hCoV-19/Portugal/PT0659a/2020\|EPI_ISL_511483\|2020- | ···················· | ···················· | ···················· | ···················· | ···················· | ···················· |
| F | 8874 hCoV-19/Portugal/PT0932/2020\|EPI_ISL_511484\|2020-0 | ···················· | ·A·················· | ···················· | ···················· | ···················· | ···················· |
| F | 8875 hCoV-19/Portugal/PT0933/2020\|EPI_ISL_511485\|2020-0 | ···················· | ···················· | ···················· | ···················· | ···················· | ···················· |
| F | 8876 hCoV-19/Portugal/PT0934/2020\|EPI_ISL_511486\|2020-0 | ···················· | ···················· | ···················· | ···················· | ···················· | ···················· |
| F | 8877 hCoV-19/Portugal/PT0936/2020\|EPI_ISL_511487\|2020-0 | ···················· | ···················· | ···················· | ···················· | ···················· | ···················· |
| F | 8878 hCoV-19/Portugal/PT0937/2020\|EPI_ISL_511488\|2020-0 | ···················· | ···················· | ···················· | ···················· | ···················· | ···················· |
| F | 8879 hCoV-19/Portugal/PT0938/2020\|EPI_ISL_511489\|2020-0 | ···················· | ···················· | ···················· | ···················· | ···················· | ···················· |
| F | 8880 hCoV-19/Portugal/PT0940/2020\|EPI_ISL_511490\|2020-0 | ···················· | ···················· | ···················· | ···················· | ···················· | ···················· |
| F | 8882 hCoV-19/Portugal/PT0942/2020\|EPI_ISL_511492\|2020-0 | ···················· | ···················· | ···················· | ···················· | ···················· | ···················· |
| F | 8883 hCoV-19/Portugal/PT0943/2020\|EPI_ISL_511493\|2020-0 | ···················· | ···················· | ···················· | ···················· | ···················· | ···················· |
| F | 8884 hCoV-19/Portugal/PT0944/2020\|EPI_ISL_511494\|2020-0 | ···················· | ···················· | ···················· | ···················· | ···················· | ···················· |
| F | 8885 hCoV-19/Portugal/PT0945/2020\|EPI_ISL_511495\|2020-0 | ···················· | ···················· | ···················· | ···················· | ···················· | ···················· |
| F | 8886 hCoV-19/Portugal/PT0946/2020\|EPI_ISL_511496\|2020-0 | ···················· | ···················· | ···················· | ···················· | ···················· | ···················· |
| F | 8887 hCoV-19/Portugal/PT0947/2020\|EPI_ISL_511497\|2020-0 | ···················· | ···················· | ···················· | ···················· | ···················· | ···················· |
| F | 8888 hCoV-19/Portugal/PT0949/2020\|EPI_ISL_511499\|2020-0 | ···················· | ···················· | ···················· | ···················· | ···················· | ···················· |
| F | 8889 hCoV-19/Australia/WA59/2020\|EPI_ISL_512712\|2020-04 | ···················· | ···················· | ···················· | ···················· | ···················· | ···················· |
| F | 8890 hCoV-19/Australia/WA60/2020\|EPI_ISL_512713\|2020-04 | ···················· | ···················· | ···················· | ···················· | ··C················· | ···················· |
| F | 8891 hCoV-19/Australia/WA61/2020\|EPI_ISL_512714\|2020-04 | ···················· | ···················· | ···················· | ···················· | ···················· | ···················· |
| F | 8892 hCoV-19/Australia/WA62/2020\|EPI_ISL_512715\|2020-05 | ···················· | ·A·················· | ···················· | ···················· | ···················· | ··A················· |
| F | 8893 hCoV-19/Australia/WA63/2020\|EPI_ISL_512716\|2020-05 | ···················· | ·A·················· | ···················· | ···················· | ···················· | ··A················· |
| F | 8894 hCoV-19/Australia/WA64/2020\|EPI_ISL_512717\|2020-05 | ···················· | ···················· | ···················· | ···················· | ···················· | ···············A···· |
| F | 8895 hCoV-19/Australia/WA66/2020\|EPI_ISL_512719\|2020-06 | ···················· | ···················· | ···················· | ···················· | ···················· | ···················· |
| F | 8896 hCoV-19/Australia/WA67/2020\|EPI_ISL_512720\|2020-06 | ···················· | ···················· | ···················· | ···················· | ···················· | ···················· |
| F | 8897 hCoV-19/Australia/WA69/2020\|EPI_ISL_512722\|2020-07 | ···················· | ·A·················· | ···················· | ···················· | ···················· | ···················· |
| F | 8898 hCoV-19/Australia/WA70/2020\|EPI_ISL_512723\|2020-07 | ···················· | ·A·················· | ···················· | ···················· | ···················· | ···················· |
| F | 8899 hCoV-19/Australia/WA71/2020\|EPI_ISL_512724\|2020-07 | ···················· | ···················· | ···················· | ···················· | ···················· | ···················· |
| F | 8900 hCoV-19/Australia/WA72/2020\|EPI_ISL_512725\|2020-07 | ···················· | ·A·················· | ···················· | ···················· | ···················· | ···················· |
| F | 8901 hCoV-19/Australia/WA74/2020\|EPI_ISL_512727\|2020-07 | ···················· | ···················· | ···················· | ···················· | ···················· | ···················· |
| F | 8902 hCoV-19/Australia/WA75/2020\|EPI_ISL_512728\|2020-07 | ···················· | ·A·················· | ···················· | ···················· | ···················· | ···················· |
| F | 8903 hCoV-19/Australia/WA77/2020\|EPI_ISL_512730\|2020-07 | ···················· | ·A·················· | ···················· | ···················· | ···················· | ···················· |
| F | 8904 hCoV-19/Australia/WA79/2020\|EPI_ISL_512732\|2020-07 | ···················· | ·A·················· | ···················· | ···················· | ···················· | ···················· |
| F | 8905 hCoV-19/Australia/WA81/2020\|EPI_ISL_512734\|2020-07 | ···················· | ·A·················· | ···················· | ···················· | ···················· | ···················· |
| F | 8906 hCoV-19/Australia/WA82/2020\|EPI_ISL_512735\|2020-07 | ···················· | ·A·················· | ···················· | ···················· | ···················· | ···················· |
| F | 8907 hCoV-19/Australia/WA84/2020\|EPI_ISL_512737\|2020-05 | ···················· | ·A·················· | ···················· | ···················· | ···················· | ··A················· |
| F | 8908 hCoV-19/Australia/WA85/2020\|EPI_ISL_512738\|2020-03 | ···················· | ···················· | ···················· | ···················· | ···················· | ···················· |
| F | 8909 hCoV-19/Australia/WA86/2020\|EPI_ISL_512739\|2020-03 | ···················· | ···················· | ···················· | ···················· | ···················· | ···················· |
| F | 8910 hCoV-19/Australia/WA88/2020\|EPI_ISL_512741\|2020-03 | ···················· | ···················· | ···················· | ···················· | ···················· | ···················· |
| F | 8911 hCoV-19/Australia/WA89/2020\|EPI_ISL_512742\|2020-03 | ···················· | ···················· | ···················· | ···················· | ···················· | ···················· |
| F | 8912 hCoV-19/Australia/WA90/2020\|EPI_ISL_512743\|2020-03 | ···················· | ···················· | ···················· | ···················· | ···················· | ···················· |
| F | 8913 hCoV-19/Australia/WA91/2020\|EPI_ISL_512744\|2020-03 | ···················· | ···················· | ···················· | ···················· | ···················· | ···················· |
| F | 8914 hCoV-19/Australia/WA97/2020\|EPI_ISL_512750\|2020-03 | ···················· | ···················· | ···················· | ···················· | ···················· | ···················· |
| F | 8915 hCoV-19/Australia/WA99/2020\|EPI_ISL_512752\|2020-04 | ···················· | ···················· | ···················· | ···················· | ···················· | ···················· |
| F | 8916 hCoV-19/Australia/WA101/2020\|EPI_ISL_512754\|2020-0 | ···················· | ···················· | ···················· | ···················· | ···················· | ···················· |
| F | 8917 hCoV-19/Australia/WA102/2020\|EPI_ISL_512755\|2020-0 | ···················· | ···················· | ···················· | ···················· | ···················· | ···················· |
| F | 8918 hCoV-19/USA/WY-USAFSAM-S015/2020\|EPI_ISL_512789\|20 | ···················· | ···················· | ···················· | ···················· | ···················· | ···················· |
| F | 8919 hCoV-19/USA/WY-USAFSAM-S016/2020\|EPI_ISL_512790\|20 | ···················· | ···················· | ···················· | ···················· | ···················· | ···················· |
| F | 8920 hCoV-19/USA/MS-USAFSAM-S021/2020\|EPI_ISL_512795\|20 | ···················· | ···················· | ···················· | ···················· | ···················· | ···················· |
| F | 8921 hCoV-19/USA/MS-USAFSAM-S023/2020\|EPI_ISL_512797\|20 | ···················· | ···················· | ···················· | ···················· | ···················· | ···················· |
| F | 8922 hCoV-19/USA/MS-USAFSAM-S025/2020\|EPI_ISL_512798\|20 | ···················· | ···················· | ···················· | ···················· | ···················· | ···················· |
| F | 8923 hCoV-19/USA/ID-USAFSAM-S028/2020\|EPI_ISL_512799\|20 | ···················· | ···················· | ···················· | ···················· | ···················· | ···················· |

| | | | | | | | |
|---|---|---|---|---|---|---|---|
| F | 8924 hCoV-19/South Korea/KCDC2597/2020\|EPI_ISL_515000\|2 | ···················· | ···················· | ···················· | ···················· | ···················· | ···················· |
| F | 8925 hCoV-19/South Korea/KCDC2598/2020\|EPI_ISL_515001\|2 | ···················· | ···················· | ···················· | ···················· | ···················· | ···················· |
| F | 8926 hCoV-19/South Korea/KCDC2599/2020\|EPI_ISL_515002\|2 | ···················· | ···················· | ···················· | ···················· | ···················· | ···················· |
| F | 8927 hCoV-19/South Korea/KCDC2600/2020\|EPI_ISL_515003\|2 | ···················· | ···················· | ···················· | ···················· | ···················· | ···················· |
| F | 8928 hCoV-19/South Korea/KCDC2601/2020\|EPI_ISL_515004\|2 | ···················· | ···················· | ···················· | ···················· | ···················· | ···················· |
| F | 8929 hCoV-19/South Korea/KCDC2602/2020\|EPI_ISL_515005\|2 | ···················· | ···················· | ···················· | ···················· | ···················· | ···················· |
| F | 8930 hCoV-19/South Korea/KCDC2603/2020\|EPI_ISL_515006\|2 | ···················· | ···················· | ···················· | ···················· | ···················· | ···················· |
| F | 8931 hCoV-19/South Korea/KCDC2604/2020\|EPI_ISL_515007\|2 | ···················· | ···················· | ···················· | ···················· | ···················· | ···················· |
| F | 8932 hCoV-19/South Korea/KCDC2605/2020\|EPI_ISL_515008\|2 | ···················· | ···················· | ···················· | ···················· | ···················· | ···················· |
| F | 8933 hCoV-19/South Korea/KCDC2606/2020\|EPI_ISL_515009\|2 | ···················· | ···················· | ···················· | ···················· | ···················· | ···················· |
| F | 8934 hCoV-19/South Korea/KCDC2607/2020\|EPI_ISL_515010\|2 | ···················· | ···················· | ···················· | ···················· | ···················· | ···················· |
| F | 8935 hCoV-19/South Korea/KCDC2608/2020\|EPI_ISL_515011\|2 | ···················· | ···················· | ···················· | ···················· | ···················· | ···················· |
| F | 8936 hCoV-19/South Korea/KCDC2609/2020\|EPI_ISL_515012\|2 | ···················· | ···················· | ···················· | ···················· | ···················· | ···················· |
| F | 8937 hCoV-19/South Korea/KCDC2610/2020\|EPI_ISL_515013\|2 | ···················· | ···················· | ···················· | ···················· | ···················· | ···················· |
| F | 8938 hCoV-19/South Korea/KCDC2611/2020\|EPI_ISL_515014\|2 | ···················· | ···················· | ···················· | ···················· | ···················· | ···················· |
| F | 8939 hCoV-19/South Korea/KCDC2612/2020\|EPI_ISL_515015\|2 | ···················· | ···················· | ···················· | ···················· | ···················· | ···················· |
| F | 8940 hCoV-19/South Korea/KCDC2613/2020\|EPI_ISL_515016\|2 | ···················· | ···················· | ···················· | ···················· | ···················· | ···················· |
| F | 8941 hCoV-19/South Korea/KCDC2614/2020\|EPI_ISL_515017\|2 | ···················· | ···················· | ···················· | ···················· | ···················· | ···················· |
| F | 8942 hCoV-19/South Korea/KCDC2615/2020\|EPI_ISL_515018\|2 | ···················· | ···················· | ···················· | ···················· | ···················· | ···················· |
| F | 8943 hCoV-19/South Korea/KCDC2616/2020\|EPI_ISL_515019\|2 | ···················· | ···················· | ···················· | ···················· | ···················· | ···················· |
| F | 8944 hCoV-19/USA/MI-MDHHS-SC21575/2020\|EPI_ISL_516302\|2 | ···················· | ···················· | ···················· | ···················· | ···················· | ···················· |
| F | 8945 hCoV-19/USA/MI-MDHHS-SC21579/2020\|EPI_ISL_516306\|2 | ···················· | ···················· | ···················· | ···················· | ···················· | ···················· |
| F | 8946 hCoV-19/USA/MI-MDHHS-SC21581/2020\|EPI_ISL_516308\|2 | ···················· | ···················· | ···················· | ···················· | ···················· | ···················· |
| F | 8947 hCoV-19/USA/MI-MDHHS-SC21584/2020\|EPI_ISL_516311\|2 | ···················· | ···················· | ···················· | ···················· | ···················· | ···················· |
| F | 8948 hCoV-19/USA/MI-MDHHS-SC21587/2020\|EPI_ISL_516314\|2 | ···················· | ···················· | ···················· | ···················· | ···················· | ···················· |
| F | 8949 hCoV-19/USA/MI-MDHHS-SC21589/2020\|EPI_ISL_516316\|2 | ···················· | ···················· | ···················· | ···················· | ···················· | ···················· |
| F | 8950 hCoV-19/USA/MI-MDHHS-SC21590/2020\|EPI_ISL_516317\|2 | ···················· | ···················· | ···················· | ···················· | ···················· | ···················· |
| F | 8951 hCoV-19/USA/MI-MDHHS-SC21594/2020\|EPI_ISL_516321\|2 | ···················· | ···················· | ···················· | ···················· | ···················· | ···················· |
| F | 8952 hCoV-19/USA/MI-MDHHS-SC21595/2020\|EPI_ISL_516322\|2 | ···················· | ···················· | ···················· | ···················· | ···················· | ···················· |
| F | 8953 hCoV-19/USA/MI-MDHHS-SC21597/2020\|EPI_ISL_516324\|2 | ···················· | ···················· | ···················· | ···················· | ···················· | ···················· |
| F | 8954 hCoV-19/USA/MI-MDHHS-SC21602/2020\|EPI_ISL_516329\|2 | ···················· | ···················· | ···················· | ···················· | ···················· | ···················· |
| F | 8955 hCoV-19/USA/MI-MDHHS-SC21603/2020\|EPI_ISL_516330\|2 | ···················· | ···················· | ···················· | ···················· | ···················· | ···················· |
| F | 8956 hCoV-19/USA/MI-MDHHS-SC21604/2020\|EPI_ISL_516331\|2 | ···················· | ···················· | ···················· | ···················· | ···················· | ···················· |
| F | 8957 hCoV-19/USA/MI-MDHHS-SC21605/2020\|EPI_ISL_516332\|2 | ···················· | ···················· | ···················· | ···················· | ···················· | ···················· |
| F | 8958 hCoV-19/USA/MI-MDHHS-SC21607/2020\|EPI_ISL_516334\|2 | ···················· | ···················· | ···················· | ···················· | ···················· | ···················· |
| F | 8959 hCoV-19/USA/MI-MDHHS-SC21608/2020\|EPI_ISL_516335\|2 | ···················· | ···················· | ···················· | ···················· | ···················· | ···················· |
| F | 8960 hCoV-19/USA/MI-MDHHS-SC21621/2020\|EPI_ISL_516348\|2 | ···················· | ···················· | ···················· | ···················· | ···················· | ···················· |
| F | 8961 hCoV-19/USA/MN-MDH-1330/2020\|EPI_ISL_496916\|2020-0 | ···················· | ·A·················· | ···················· | ···················· | ···················· | ···················· |
| F | 8962 hCoV-19/USA/MN-MDH-1331/2020\|EPI_ISL_496917\|2020-0 | ···················· | ···················· | ···················· | ···················· | ···················· | ···················· |
| F | 8963 hCoV-19/USA/MN-MDH-1332/2020\|EPI_ISL_496918\|2020-0 | ···················· | ···················· | ···················· | ···················· | ···················· | ···················· |
| F | 8964 hCoV-19/USA/MN-MDH-1333/2020\|EPI_ISL_496919\|2020-0 | ···················· | ···················· | ···················· | ···················· | ···················· | ···················· |
| F | 8965 hCoV-19/USA/MN-MDH-1334/2020\|EPI_ISL_496920\|2020-0 | ···················· | ···················· | ···················· | ···················· | ···················· | ···················· |
| F | 8966 hCoV-19/USA/MN-MDH-1335/2020\|EPI_ISL_496921\|2020-0 | ···················· | ···················· | ···················· | ···················· | ···················· | ···················· |
| F | 8967 hCoV-19/USA/MN-MDH-1336/2020\|EPI_ISL_496922\|2020-0 | ···················· | ···················· | ···················· | ···················· | ···················· | ···················· |
| F | 8968 hCoV-19/USA/MN-MDH-1337/2020\|EPI_ISL_496923\|2020-0 | ···················· | ···················· | ···················· | ···················· | ···················· | ···················· |
| F | 8969 hCoV-19/USA/MN-MDH-1338/2020\|EPI_ISL_496924\|2020-0 | ···················· | ···················· | ···················· | ···················· | ···················· | ···················· |
| F | 8970 hCoV-19/USA/MN-MDH-1339/2020\|EPI_ISL_496925\|2020-0 | ···················· | ···················· | ···················· | ···················· | ···················· | ···················· |
| F | 8971 hCoV-19/USA/MN-MDH-1340/2020\|EPI_ISL_496926\|2020-0 | ···················· | ···················· | ···················· | ···················· | ···················· | ···················· |
| F | 8972 hCoV-19/USA/MN-MDH-1341/2020\|EPI_ISL_496927\|2020-0 | ···················· | ···················· | ···················· | ···················· | ···················· | ···················· |
| F | 8973 hCoV-19/USA/MN-MDH-1342/2020\|EPI_ISL_496928\|2020-0 | ···················· | ···················· | ···················· | ···················· | ···················· | ···················· |
| F | 8974 hCoV-19/USA/MN-MDH-1343/2020\|EPI_ISL_496929\|2020-0 | ···················· | ···················· | ···················· | ···················· | ···················· | ···················· |
| F | 8975 hCoV-19/USA/MN-MDH-1344/2020\|EPI_ISL_496930\|2020-0 | ···················· | ···················· | ···················· | ···················· | ···················· | ···················· |
| F | 8976 hCoV-19/USA/WA-S2299/2020\|EPI_ISL_496931\|2020-06-1 | ···················· | ···················· | ···················· | ···················· | ···················· | ···················· |
| F | 8977 hCoV-19/USA/WA-S2300/2020\|EPI_ISL_496932\|2020-06-1 | ···················· | ···················· | ···················· | ···················· | ···················· | ···················· |
| F | 8978 hCoV-19/USA/WA-S2301/2020\|EPI_ISL_496933\|2020-06-1 | ···················· | ···················· | ···················· | ···················· | ···················· | ···················· |
| F | 8979 hCoV-19/USA/WA-S2302/2020\|EPI_ISL_496934\|2020-06-1 | ···················· | ···················· | ···················· | ···················· | ···················· | ···················· |
| F | 8980 hCoV-19/USA/WA-S2303/2020\|EPI_ISL_496935\|2020-06-1 | ···················· | ···················· | ···················· | ···················· | ···················· | ···················· |
| F | 8981 hCoV-19/USA/WA-S2304/2020\|EPI_ISL_496936\|2020-06-1 | ···················· | ···················· | ···················· | ···················· | ···················· | ···················· |
| F | 8982 hCoV-19/USA/WA-S2305/2020\|EPI_ISL_496937\|2020-06-1 | ···················· | ···················· | ···················· | ···················· | ···················· | ···················· |
| F | 8983 hCoV-19/USA/WA-S2306/2020\|EPI_ISL_496938\|2020-06-1 | ···················· | ···················· | ···················· | ···················· | ···················· | ···················· |
| F | 8984 hCoV-19/USA/WA-S2307/2020\|EPI_ISL_496939\|2020-06-1 | ···················· | ···················· | ···················· | ···················· | ···················· | ···················· |
| F | 8985 hCoV-19/USA/WA-S2308/2020\|EPI_ISL_496940\|2020-06-1 | ···················· | ···················· | ···················· | ···················· | ···················· | ···················· |
| F | 8986 hCoV-19/USA/WA-S2309/2020\|EPI_ISL_496941\|2020-06-1 | ···················· | ···················· | ···················· | ···················· | ···················· | ···················· |
| F | 8987 hCoV-19/USA/WA-S2310/2020\|EPI_ISL_496942\|2020-06-1 | ···················· | ·A·················· | ···················· | ···················· | ···················· | ···················· |
| F | 8988 hCoV-19/USA/WA-S2311/2020\|EPI_ISL_496943\|2020-06-1 | ···················· | ···················· | ···················· | ···················· | ···················· | ···················· |
| F | 8989 hCoV-19/USA/WA-S2313/2020\|EPI_ISL_496945\|2020-06-1 | ···················· | ····A··············· | ···················· | ···················· | ···················· | ···················· |
| F | 8990 hCoV-19/USA/WA-S2315/2020\|EPI_ISL_496947\|2020-06-1 | ···················· | ···················· | ···················· | ···················· | ···················· | ···················· |
| F | 8991 hCoV-19/USA/WA-S2316/2020\|EPI_ISL_496948\|2020-06-1 | ···················· | ···················· | ···················· | ···················· | ···················· | ···················· |
| F | 8992 hCoV-19/USA/WA-S2317/2020\|EPI_ISL_496949\|2020-06-1 | ···················· | ···················· | ···················· | ···················· | ···················· | ···················· |
| F | 8993 hCoV-19/USA/WA-S2321/2020\|EPI_ISL_496953\|2020-06-1 | ···················· | ···················· | ···················· | ···················· | ···················· | ···················· |
| F | 8994 hCoV-19/USA/WA-S2322/2020\|EPI_ISL_496954\|2020-06-1 | ···················· | ···················· | ···················· | ···················· | ···················· | ···················· |
| F | 8995 hCoV-19/USA/WA-S2323/2020\|EPI_ISL_496955\|2020-06-1 | ···················· | ···················· | ···················· | ···················· | ···················· | ···················· |
| F | 8996 hCoV-19/USA/WA-S2325/2020\|EPI_ISL_496957\|2020-06-1 | ···················· | ···················· | ···················· | ···················· | ···················· | ···················· |
| F | 8997 hCoV-19/USA/WA-S2326/2020\|EPI_ISL_496958\|2020-06-1 | ···················· | ···················· | ···················· | ···················· | ···················· | ···················· |
| F | 8998 hCoV-19/USA/WA-S2340/2020\|EPI_ISL_496972\|2020-06-1 | ···················· | ····A··············· | ···················· | ···················· | ···················· | ···················· |
| F | 8999 hCoV-19/USA/WA-S2342/2020\|EPI_ISL_496974\|2020-06-1 | ···················· | ···················· | ···················· | ···················· | ···················· | ···················· |
| F | 9000 hCoV-19/USA/WA-S2343/2020\|EPI_ISL_496975\|2020-06-1 | ···················· | ···················· | ···················· | ···················· | ···················· | ···················· |
| F | 9001 hCoV-19/USA/WA-S2344/2020\|EPI_ISL_496976\|2020-06-1 | ···················· | ···················· | ···················· | ···················· | ···················· | ···················· |
| F | 9002 hCoV-19/USA/WA-S2345/2020\|EPI_ISL_496977\|2020-06-1 | ···················· | ···················· | ···················· | ···················· | ···················· | ···················· |
| F | 9003 hCoV-19/USA/WA-S2346/2020\|EPI_ISL_496978\|2020-06-1 | ···················· | ···················· | ···················· | ···················· | ···················· | ···················· |

| | | | | | | | | |
|---|---|---|---|---|---|---|---|---|
| F | 9004 | hCoV-19/USA/WA-S2347/2020\|EPI_ISL_496979\|2020-06-1 | .................... | .................... | ..................... | .................... | .................... | .................... |
| F | 9005 | hCoV-19/USA/WA-S2348/2020\|EPI_ISL_496980\|2020-06-1 | .................... | .................... | ..................... | .................... | .................... | .................... |
| F | 9006 | hCoV-19/USA/WA-S2349/2020\|EPI_ISL_496981\|2020-06-1 | .................... | .................... | ..................... | .................... | .................... | .................... |
| F | 9007 | hCoV-19/USA/WA-S2350/2020\|EPI_ISL_496982\|2020-06-1 | .................... | .................... | ..................... | .................... | .................... | .................... |
| F | 9008 | hCoV-19/USA/WA-S2351/2020\|EPI_ISL_496983\|2020-06-1 | .................... | .................... | ..................... | .................... | .................... | .................... |
| F | 9009 | hCoV-19/USA/WA-S2352/2020\|EPI_ISL_496984\|2020-06-1 | .................... | .................... | ..................... | .................... | .................... | .................... |
| F | 9010 | hCoV-19/USA/WA-S2353/2020\|EPI_ISL_496985\|2020-06-1 | .................... | .................... | ..................... | .................... | .................... | .................... |
| F | 9011 | hCoV-19/USA/WA-S2355/2020\|EPI_ISL_496987\|2020-06-1 | .................... | .................... | ..................... | .................... | .................... | .................... |
| F | 9012 | hCoV-19/USA/WA-S2356/2020\|EPI_ISL_496988\|2020-06-1 | .................... | .................... | ..................... | .................... | .................... | .................... |
| F | 9013 | hCoV-19/USA/WA-S2357/2020\|EPI_ISL_496989\|2020-06-1 | .................... | .................... | ..................... | .................... | .................... | .................... |
| F | 9014 | hCoV-19/USA/WA-S2358/2020\|EPI_ISL_496990\|2020-06-1 | .................... | .................... | ..................... | .................... | .................... | .................... |
| F | 9015 | hCoV-19/USA/WA-S2359/2020\|EPI_ISL_496991\|2020-06-1 | .................... | .................... | ..................... | .................... | .................... | .................... |
| F | 9016 | hCoV-19/USA/WA-S2360/2020\|EPI_ISL_496992\|2020-06-1 | .................... | .................... | ..................... | .................... | .................... | .................... |
| F | 9017 | hCoV-19/South Korea/KCDC2116/2020\|EPI_ISL_498017\|2 | .................... | .................... | ..................... | .................... | .................... | .................... |
| F | 9018 | hCoV-19/South Korea/KCDC2117/2020\|EPI_ISL_498018\|2 | .................... | .................... | ..................... | .................... | .................... | .................... |
| F | 9019 | hCoV-19/South Korea/KCDC2118/2020\|EPI_ISL_498019\|2 | .................... | .................... | ..................... | .................... | .................... | .................... |
| F | 9020 | hCoV-19/South Korea/KCDC2126/2020\|EPI_ISL_498027\|2 | .................... | .................... | ..................... | .................... | .................... | .................... |
| F | 9021 | hCoV-19/South Korea/KCDC2127/2020\|EPI_ISL_498028\|2 | .................... | .................... | ..................... | .................... | .................... | .................... |
| F | 9022 | hCoV-19/South Korea/KCDC2128/2020\|EPI_ISL_498029\|2 | .................... | .................... | ..................... | .................... | .................... | .................... |
| F | 9023 | hCoV-19/South Korea/KCDC2129/2020\|EPI_ISL_498030\|2 | .................... | .................... | ..................... | .................... | .................... | .................... |
| F | 9024 | hCoV-19/South Korea/KCDC2130/2020\|EPI_ISL_498031\|2 | .................... | .................... | ..................... | .................... | .................... | .................... |
| F | 9025 | hCoV-19/South Korea/KCDC2131/2020\|EPI_ISL_498032\|2 | .................... | .................... | ..................... | .................... | .................... | .................... |
| F | 9026 | hCoV-19/South Korea/KCDC2132/2020\|EPI_ISL_498033\|2 | .................... | .................... | ..................... | .................... | .................... | .................... |
| F | 9027 | hCoV-19/South Korea/KCDC2133/2020\|EPI_ISL_498034\|2 | .................... | .................... | ..................... | .................... | .................... | .................... |
| F | 9028 | hCoV-19/South Korea/KCDC2134/2020\|EPI_ISL_498035\|2 | .................... | .................... | ..................... | .................... | .................... | .................... |
| F | 9029 | hCoV-19/South Korea/KCDC2135/2020\|EPI_ISL_498036\|2 | .................... | .................... | ..................... | .................... | .................... | .................... |
| F | 9030 | hCoV-19/South Korea/KCDC2136/2020\|EPI_ISL_498037\|2 | .................... | .................... | ..................... | .................... | .................... | .................... |
| F | 9031 | hCoV-19/Wales/PHWC-168E74/2020\|EPI_ISL_499358\|2020 | .................... | .................... | ..................... | .................... | .................... | .................... |
| F | 9032 | hCoV-19/Wales/PHWC-168EA1/2020\|EPI_ISL_499360\|2020 | .................... | .................... | ..................... | .................... | .................... | .................... |
| F | 9033 | hCoV-19/Wales/PHWC-168EB0/2020\|EPI_ISL_499361\|2020 | .................... | .................... | ..................... | .................... | .................... | .................... |
| F | 9034 | hCoV-19/Wales/PHWC-168ECF/2020\|EPI_ISL_499362\|2020 | .................... | .................... | ..................... | .................... | .................... | .................... |
| F | 9035 | hCoV-19/Wales/PHWC-168EDE/2020\|EPI_ISL_499363\|2020 | .................... | .................... | ..................... | .................... | .................... | .................... |
| F | 9036 | hCoV-19/Wales/PHWC-168EED/2020\|EPI_ISL_499364\|2020 | .................... | .................... | ..................... | .................... | .................... | .................... |
| F | 9037 | hCoV-19/Wales/PHWC-168F08/2020\|EPI_ISL_499366\|2020 | .................... | .................... | ..................... | .................... | .................... | .................... |
| F | 9038 | hCoV-19/Wales/PHWC-168F17/2020\|EPI_ISL_499367\|2020 | .................... | .................... | ..................... | .................... | .................... | .................... |
| F | 9039 | hCoV-19/Wales/PHWC-168F26/2020\|EPI_ISL_499368\|2020 | .................... | .................... | ..................... | .................... | .................... | .................... |
| F | 9040 | hCoV-19/Australia/NSW2212/2020\|EPI_ISL_500667\|2020 | .................... | .................... | ..................... | .................... | .................... | .................... |
| F | 9041 | hCoV-19/Australia/NSW2214/2020\|EPI_ISL_500668\|2020 | .................... | .................... | ..................... | .................... | .................... | .................... |
| F | 9042 | hCoV-19/Australia/NSW2216/2020\|EPI_ISL_500669\|2020 | .................... | .................... | ..................... | .................... | .................... | .................... |
| F | 9043 | hCoV-19/Australia/NSW2218/2020\|EPI_ISL_500670\|2020 | .................... | .................... | ..................... | .................... | .................... | .................... |
| F | 9044 | hCoV-19/Australia/NSW2220/2020\|EPI_ISL_500671\|2020 | .................... | .................... | ..................... | .................... | .................... | .................... |
| F | 9045 | hCoV-19/Australia/NSW2221/2020\|EPI_ISL_500672\|2020 | .................... | .................... | ..................... | .................... | .................... | .................... |
| F | 9046 | hCoV-19/Australia/NSW2222/2020\|EPI_ISL_500673\|2020 | .................... | .................... | ..................... | .................... | .................... | .................... |
| F | 9047 | hCoV-19/Australia/NSW2226/2020\|EPI_ISL_500674\|2020 | .................... | .................... | ..................... | .................... | .................... | .................... |
| F | 9048 | hCoV-19/Australia/NSW2227/2020\|EPI_ISL_500675\|2020 | .................... | .................... | ..................... | .................... | .................... | .................... |
| F | 9049 | hCoV-19/Australia/NSW2228/2020\|EPI_ISL_500676\|2020 | .................... | .................... | ..................... | .................... | .................... | .................... |
| F | 9050 | hCoV-19/Australia/NSW2229/2020\|EPI_ISL_500677\|2020 | .................... | .................... | ..................... | .................... | .................... | .................... |
| F | 9051 | hCoV-19/Australia/NSW2238/2020\|EPI_ISL_500680\|2020 | .................... | .................... | ..................... | .................... | .................... | .................... |
| F | 9052 | hCoV-19/USA/VA-DCLS-0573/2020\|EPI_ISL_500831\|2020- | .................... | .................... | ..................... | .................... | .................... | .................... |
| F | 9053 | hCoV-19/USA/VA-DCLS-0574/2020\|EPI_ISL_500832\|2020- | .................... | .................... | ..................... | .................... | .................... | .................... |
| F | 9054 | hCoV-19/USA/VA-DCLS-0670/2020\|EPI_ISL_500833\|2020- | .................... | .................... | ..................... | .................... | .................... | .................... |
| F | 9055 | hCoV-19/USA/VA-DCLS-0671/2020\|EPI_ISL_500834\|2020- | .................... | .................... | ..................... | .................... | .................... | .................... |
| F | 9056 | hCoV-19/USA/VA-DCLS-0672/2020\|EPI_ISL_500835\|2020- | .................... | .................... | ..................... | .................... | .................... | .................... |
| F | 9057 | hCoV-19/USA/VA-DCLS-0673/2020\|EPI_ISL_500836\|2020- | .................... | .................... | ..................... | .................... | .................... | .................... |
| F | 9058 | hCoV-19/USA/VA-DCLS-0675/2020\|EPI_ISL_500838\|2020- | .................... | .................... | ..................... | .................... | .................... | .................... |
| F | 9059 | hCoV-19/USA/VA-DCLS-0676/2020\|EPI_ISL_500839\|2020- | .................... | .................... | ..................... | .................... | .................... | .................... |
| F | 9060 | hCoV-19/USA/VA-DCLS-0677/2020\|EPI_ISL_500840\|2020- | .................... | .................... | ..................... | .................... | .................... | .................... |
| F | 9061 | hCoV-19/USA/VA-DCLS-0678/2020\|EPI_ISL_500841\|2020- | .................... | .................... | ..................... | .................... | .................... | .................... |
| F | 9062 | hCoV-19/USA/VA-DCLS-0679/2020\|EPI_ISL_500842\|2020- | .................... | .................... | ..................... | .................... | .................... | .................... |
| F | 9063 | hCoV-19/South Korea/KCDC2154/2020\|EPI_ISL_506948\|2 | .................... | .................... | ..................... | .................... | .................... | .................... |
| F | 9064 | hCoV-19/South Korea/KCDC2155/2020\|EPI_ISL_506949\|2 | .................... | .................... | ..................... | .................... | .................... | .................... |
| F | 9065 | hCoV-19/South Korea/KCDC2156/2020\|EPI_ISL_506950\|2 | .................... | .................... | ..................... | .................... | .................... | .................... |
| F | 9066 | hCoV-19/South Korea/KCDC2157/2020\|EPI_ISL_506951\|2 | .................... | .................... | ..................... | .................... | .................... | .................... |
| F | 9067 | hCoV-19/South Korea/KCDC2158/2020\|EPI_ISL_506952\|2 | .................... | .................... | ..................... | .................... | .................... | .................... |
| F | 9068 | hCoV-19/South Korea/KCDC2159/2020\|EPI_ISL_506953\|2 | .................... | .................... | ..................... | .................... | .................... | .................... |
| F | 9069 | hCoV-19/South Korea/KCDC2160/2020\|EPI_ISL_506954\|2 | .................... | .................... | ..................... | .................... | .................... | .................... |
| F | 9070 | hCoV-19/South Korea/KCDC2161/2020\|EPI_ISL_506955\|2 | .................... | .................... | ..................... | .................... | .................... | .................... |
| F | 9071 | hCoV-19/South Korea/KCDC2162/2020\|EPI_ISL_506956\|2 | .................... | .................... | ..................... | .................... | .................... | .................... |
| F | 9072 | hCoV-19/South Korea/KCDC2163/2020\|EPI_ISL_506957\|2 | .................... | .................... | ..................... | .................... | .................... | .................... |
| F | 9073 | hCoV-19/South Korea/KCDC2164/2020\|EPI_ISL_506958\|2 | .................... | .................... | ..................... | .................... | .................... | .................... |
| F | 9074 | hCoV-19/South Korea/KCDC2165/2020\|EPI_ISL_506959\|2 | .................... | .................... | ..................... | .................... | .................... | .................... |
| F | 9075 | hCoV-19/South Korea/KCDC2166/2020\|EPI_ISL_506960\|2 | .................... | .................... | ..................... | .................... | .................... | .................... |
| F | 9076 | hCoV-19/South Korea/KCDC2167/2020\|EPI_ISL_506961\|2 | .................... | .................... | ..................... | .................... | .................... | .................... |
| F | 9077 | hCoV-19/South Korea/KCDC2168/2020\|EPI_ISL_506962\|2 | .................... | .................... | ..................... | .................... | .................... | .................... |
| F | 9078 | hCoV-19/South Korea/KCDC2169/2020\|EPI_ISL_506963\|2 | .................... | .................... | ..................... | .................... | .................... | .................... |
| F | 9079 | hCoV-19/South Korea/KCDC2170/2020\|EPI_ISL_506964\|2 | .................... | .................... | ..................... | .................... | .................... | .................... |
| F | 9080 | hCoV-19/South Korea/KCDC2171/2020\|EPI_ISL_506965\|2 | .................... | .................... | ..................... | .................... | .................... | .................... |
| F | 9081 | hCoV-19/South Korea/KCDC2172/2020\|EPI_ISL_506966\|2 | .................... | .................... | ..................... | .................... | .................... | .................... |
| F | 9082 | hCoV-19/South Korea/KCDC2173/2020\|EPI_ISL_506967\|2 | .................... | .................... | ..................... | .................... | .................... | .................... |
| F | 9083 | hCoV-19/South Korea/KCDC2174/2020\|EPI_ISL_506968\|2 | .................... | .................... | ..................... | .................... | .................... | .................... |

| | | | | | | | |
|---|---|---|---|---|---|---|---|
| F | 9084 hCoV-19/South Korea/KCDC2175/2020\|EPI_ISL_506969\|2 | .................... | .................... | .................... | .................... | .................... | .................... |
| F | 9085 hCoV-19/South Korea/KCDC2186/2020\|EPI_ISL_506980\|2 | .................... | .................... | .................... | .................... | .................... | .................... |
| F | 9086 hCoV-19/South Korea/KCDC2187/2020\|EPI_ISL_506981\|2 | .................... | .................... | .................... | .................... | .................... | .................... |
| F | 9087 hCoV-19/South Korea/KCDC2188/2020\|EPI_ISL_506982\|2 | .................... | .................... | .................... | .................... | .................... | .................... |
| F | 9088 hCoV-19/South Korea/KCDC2189/2020\|EPI_ISL_506983\|2 | .................... | .................... | .................... | .................... | .................... | .................... |
| F | 9089 hCoV-19/France/ARA-10821/2020\|EPI_ISL_509000\|2020- | .................... | .................... | .................... | .................... | .................... | .................... |
| F | 9090 hCoV-19/France/ARA-10863/2020\|EPI_ISL_509001\|2020- | .................... | .................... | .................... | .................... | .................... | .................... |
| F | 9091 hCoV-19/France/ARA-1125/2020\|EPI_ISL_509002\|2020-0 | .................... | .................... | .................... | .................... | .................... | .................... |
| F | 9092 hCoV-19/France/ARA-9584/2020\|EPI_ISL_509003\|2020-0 | .................... | .................... | .................... | .................... | .................... | .................... |
| F | 9093 hCoV-19/France/ARA-9476/2020\|EPI_ISL_509004\|2020-0 | .................... | .................... | .................... | .................... | .................... | .................... |
| F | 9094 hCoV-19/France/ARA-12151/2020\|EPI_ISL_509005\|2020- | .................... | .................... | .................... | .................... | .................... | .................... |
| F | 9095 hCoV-19/France/ARA-12197/2020\|EPI_ISL_509006\|2020- | .................... | .................... | .................... | .................... | .................... | .................... |
| F | 9096 hCoV-19/France/ARA-1276/2020\|EPI_ISL_509007\|2020-0 | .................... | .................... | .................... | .................... | .................... | .................... |
| F | 9097 hCoV-19/France/ARA-12740/2020\|EPI_ISL_509008\|2020- | .................... | .................... | .................... | .................... | .................... | .................... |
| F | 9098 hCoV-19/France/ARA-16647/2020\|EPI_ISL_509009\|2020- | .................... | .................... | .................... | .................... | .................... | .................... |
| F | 9099 hCoV-19/Poland/D_348r/2020\|EPI_ISL_509019\|2020-06 | .................... | .................... | .................... | .................... | .................... | .................... |
| F | 9100 hCoV-19/Portugal/PT1367/2020\|EPI_ISL_511326\|2020-0 | .................... | .................... | .................... | .................... | .................... | .................... |
| F | 9101 hCoV-19/Portugal/PT1368/2020\|EPI_ISL_511327\|2020-0 | .................... | .................... | .................... | .................... | .................... | .................... |
| F | 9102 hCoV-19/Portugal/PT1369/2020\|EPI_ISL_511328\|2020-0 | .................... | .................... | .................... | .................... | .................... | .................... |
| F | 9103 hCoV-19/Portugal/PT1372/2020\|EPI_ISL_511331\|2020-0 | .................... | .................... | .................... | .................... | .................... | .................... |
| F | 9104 hCoV-19/Portugal/PT1373/2020\|EPI_ISL_511332\|2020-0 | .................... | .................... | .................... | .................... | .................... | .................... |
| F | 9105 hCoV-19/Portugal/PT1374/2020\|EPI_ISL_511333\|2020-0 | .................... | .................... | .................... | .................... | .................... | .................... |
| F | 9106 hCoV-19/Portugal/PT1375/2020\|EPI_ISL_511334\|2020-0 | .................... | .................... | .................... | .................... | .................... | .................... |
| F | 9107 hCoV-19/Portugal/PT1377/2020\|EPI_ISL_511336\|2020-0 | .................... | .................... | .................... | .................... | .................... | .................... |
| F | 9108 hCoV-19/Costa Rica/INC-0051/2020\|EPI_ISL_512657\|20 | .................... | .................... | .................... | .................... | .................... | .................... |
| F | 9109 hCoV-19/Costa Rica/INC-0052/2020\|EPI_ISL_512658\|20 | .................... | .................... | .................... | .................... | .................... | .................... |
| F | 9110 hCoV-19/Costa Rica/INC-0053/2020\|EPI_ISL_512659\|20 | .................... | .................... | .................... | .................... | .................... | .................... |
| F | 9111 hCoV-19/Costa Rica/INC-0054/2020\|EPI_ISL_512660\|20 | .................... | .................... | .................... | .................... | .................... | .................... |
| F | 9112 hCoV-19/Costa Rica/INC-0055/2020\|EPI_ISL_512661\|20 | .................... | .................... | .................... | .................... | .................... | .................... |
| F | 9113 hCoV-19/Costa Rica/INC-0056/2020\|EPI_ISL_512662\|20 | .................... | .................... | .................... | .................... | .................... | .................... |
| F | 9114 hCoV-19/Costa Rica/INC-0057/2020\|EPI_ISL_512663\|20 | .................... | .................... | .................... | .................... | .................... | .................... |
| F | 9115 hCoV-19/Costa Rica/INC-0058/2020\|EPI_ISL_512664\|20 | .................... | .................... | .................... | .................... | .................... | .................... |
| F | 9116 hCoV-19/Costa Rica/INC-0059/2020\|EPI_ISL_512665\|20 | .................... | .................... | .................... | .................... | .................... | .................... |
| F | 9117 hCoV-19/Costa Rica/INC-0060/2020\|EPI_ISL_512666\|20 | .................... | .................... | .................... | .................... | .................... | .................... |
| F | 9118 hCoV-19/Costa Rica/INC-0061/2020\|EPI_ISL_512667\|20 | .................... | .................... | .................... | .................... | .................... | .................... |
| F | 9119 hCoV-19/USA/ID-USAFSAM-S029/2020\|EPI_ISL_512800\|20 | .................... | .................... | .................... | .................... | .................... | .................... |
| F | 9120 hCoV-19/USA/FL-USAFSAM-S033/2020\|EPI_ISL_512801\|20 | .................... | .................... | .................... | .................... | .................... | .................... |
| F | 9121 hCoV-19/USA/FL-USAFSAM-S034/2020\|EPI_ISL_512802\|20 | .................... | .................... | .................... | .................... | .................... | .................... |
| F | 9122 hCoV-19/USA/FL-USAFSAM-S035/2020\|EPI_ISL_512803\|20 | .................... | .................... | .................... | .................... | .................... | .................... |
| F | 9123 hCoV-19/USA/CO-USAFSAM-S040/2020\|EPI_ISL_512805\|20 | .................... | .................... | .................... | .................... | .................... | .................... |
| F | 9124 hCoV-19/USA/OH-USAFSAM-S041/2020\|EPI_ISL_512806\|20 | .................... | .................... | .................... | .................... | .................... | .................... |
| F | 9125 hCoV-19/USA/OH-USAFSAM-S042/2020\|EPI_ISL_512807\|20 | .................... | .................... | .................... | .................... | .................... | .................... |
| F | 9126 hCoV-19/USA/OH-USAFSAM-S043/2020\|EPI_ISL_512808\|20 | .................... | .................... | .................... | .................... | .................... | .................... |
| F | 9127 hCoV-19/USA/OH-USAFSAM-S044/2020\|EPI_ISL_512809\|20 | .................... | .................... | .................... | .................... | .................... | .................... |
| F | 9128 hCoV-19/Ukraine/203100362/2020\|EPI_ISL_512810\|2020 | .................... | .................... | .................... | .....-.............. | .................... | .................... |
| F | 9129 hCoV-19/Sierra Leone/KGH-G-8548/2020\|EPI_ISL_51281 | .................... | .................... | .................... | .................... | .................... | .................... |
| F | 9130 hCoV-19/Sierra Leone/KGH-G-8549/2020\|EPI_ISL_51281 | .................... | .................... | .................... | .................... | .................... | .................... |
| F | 9131 hCoV-19/Sierra Leone/KGH-G-8603/2020\|EPI_ISL_51281 | .................... | .................... | .................... | .................... | .................... | .................... |
| F | 9132 hCoV-19/Sierra Leone/KGH-G-8604/2020\|EPI_ISL_51281 | .................... | .................... | .................... | .................... | .................... | .................... |
| F | 9133 hCoV-19/Sierra Leone/KGH-G-8607/2020\|EPI_ISL_51281 | .................... | .................... | .................... | .................... | .................... | .................... |
| F | 9134 hCoV-19/Sierra Leone/KGH-G-8625/2020\|EPI_ISL_51281 | .................... | .................... | .................... | .................... | .................... | .................... |
| F | 9135 hCoV-19/Sierra Leone/KGH-G-8626/2020\|EPI_ISL_51281 | .................... | .................... | .................... | .................... | .................... | .................... |
| F | 9136 hCoV-19/Sierra Leone/KGH-G-8627/2020\|EPI_ISL_51281 | .................... | .................... | .................... | .................... | .................... | .................... |
| F | 9137 hCoV-19/Sierra Leone/KGH-G-8669/2020\|EPI_ISL_51282 | .................... | .................... | .................... | .................... | .................... | .................... |
| F | 9138 hCoV-19/Sierra Leone/KGH-G-8790/2020\|EPI_ISL_51282 | .................... | .................... | .................... | .................... | .................... | .................... |
| F | 9139 hCoV-19/South Korea/KCDC2617/2020\|EPI_ISL_515020\|2 | .................... | .................... | .................... | .................... | .................... | .................... |
| F | 9140 hCoV-19/South Korea/KCDC2618/2020\|EPI_ISL_515021\|2 | .................... | .................... | .................... | .................... | .................... | .................... |
| F | 9141 hCoV-19/South Korea/KCDC2619/2020\|EPI_ISL_515022\|2 | .................... | .................... | .................... | .................... | .................... | .................... |
| F | 9142 hCoV-19/South Korea/KCDC2620/2020\|EPI_ISL_515023\|2 | .................... | .................... | .................... | .................... | .................... | .................... |
| F | 9143 hCoV-19/South Korea/KCDC2621/2020\|EPI_ISL_515024\|2 | .................... | .................... | .................... | .................... | .................... | .................... |
| F | 9144 hCoV-19/South Korea/KCDC2622/2020\|EPI_ISL_515025\|2 | .................... | .................... | .................... | .................... | .................... | .................... |
| F | 9145 hCoV-19/South Korea/KCDC2623/2020\|EPI_ISL_515026\|2 | .................... | .................... | .................... | .................... | .................... | .................... |
| F | 9146 hCoV-19/South Korea/KCDC2624/2020\|EPI_ISL_515027\|2 | .................... | .................... | .................... | .................... | .................... | .................... |
| F | 9147 hCoV-19/South Korea/KCDC2626/2020\|EPI_ISL_515028\|2 | .................... | .................... | .................... | .................... | .................... | .................... |
| F | 9148 hCoV-19/South Korea/KCDC2627/2020\|EPI_ISL_515029\|2 | .................... | .................... | .................... | .................... | .................... | .................... |
| F | 9149 hCoV-19/South Korea/KCDC2628/2020\|EPI_ISL_515030\|2 | .................... | .................... | .................... | .................... | .................... | .................... |
| F | 9150 hCoV-19/South Korea/KCDC2629/2020\|EPI_ISL_515031\|2 | .................... | .................... | .................... | .................... | .................... | .................... |
| F | 9151 hCoV-19/South Korea/KCDC2630/2020\|EPI_ISL_515032\|2 | .................... | .................... | .................... | .................... | .................... | .................... |
| F | 9152 hCoV-19/South Korea/KCDC2631/2020\|EPI_ISL_515033\|2 | .................... | .................... | .................... | .................... | .................... | .................... |
| F | 9153 hCoV-19/South Korea/KCDC2632/2020\|EPI_ISL_515034\|2 | .................... | .................... | .................... | .................... | .................... | .................... |
| F | 9154 hCoV-19/South Korea/KCDC2633/2020\|EPI_ISL_515035\|2 | .................... | .................... | .................... | .................... | .................... | .................... |
| F | 9155 hCoV-19/South Korea/KCDC2634/2020\|EPI_ISL_515036\|2 | .................... | .................... | .................... | .................... | .................... | .................... |
| F | 9156 hCoV-19/South Korea/KCDC2635/2020\|EPI_ISL_515037\|2 | .................... | .................... | .................... | .................... | .................... | .................... |
| F | 9157 hCoV-19/South Korea/KCDC2636/2020\|EPI_ISL_515038\|2 | .................... | .................... | .................... | .................... | .................... | .................... |
| F | 9158 hCoV-19/South Korea/KCDC2637/2020\|EPI_ISL_515039\|2 | .................... | .................... | .................... | .................... | .................... | .................... |
| F | 9159 hCoV-19/South Korea/KCDC2638/2020\|EPI_ISL_515040\|2 | .................... | .................... | .................... | .................... | .................... | .................... |
| F | 9160 hCoV-19/South Korea/KCDC2639/2020\|EPI_ISL_515041\|2 | .................... | .................... | .................... | .................... | .................... | .................... |
| F | 9161 hCoV-19/South Korea/KCDC2640/2020\|EPI_ISL_515042\|2 | .................... | .................... | .................... | .................... | .................... | .................... |
| F | 9162 hCoV-19/South Korea/KCDC2641/2020\|EPI_ISL_515043\|2 | .................... | .................... | .................... | .................... | .................... | .................... |
| F | 9163 hCoV-19/South Korea/KCDC2642/2020\|EPI_ISL_515044\|2 | .................... | .................... | .................... | .................... | .................... | .................... |

| | | | | | | | |
|---|---|---|---|---|---|---|---|
| F | 9164 hCoV-19/South Korea/KCDC2643/2020\|EPI_ISL_515045\|2 | ·················· | ·················· | ·················· | ·················· | ·················· | ·················· |
| F | 9165 hCoV-19/South Korea/KCDC2644/2020\|EPI_ISL_515046\|2 | ·················· | ·················· | ·················· | ·················· | ·················· | ·················· |
| F | 9166 hCoV-19/South Korea/KCDC2645/2020\|EPI_ISL_515047\|2 | ·················· | ·················· | ·················· | ·················· | ·················· | ·················· |
| F | 9167 hCoV-19/South Korea/KCDC2646/2020\|EPI_ISL_515048\|2 | ·················· | ·················· | ·················· | ·················· | ·················· | ·················· |
| F | 9168 hCoV-19/South Korea/KCDC2647/2020\|EPI_ISL_515049\|2 | ·················· | ·················· | ·················· | ·················· | ·················· | ·················· |
| F | 9169 hCoV-19/South Korea/KCDC2648/2020\|EPI_ISL_515050\|2 | ·················· | ·················· | ·················· | ·················· | ·················· | ·················· |
| F | 9170 hCoV-19/South Korea/KCDC2650/2020\|EPI_ISL_515052\|2 | ·················· | ·················· | ·················· | ·················· | ·················· | ·················· |
| F | 9171 hCoV-19/South Korea/KCDC2651/2020\|EPI_ISL_515053\|2 | ·················· | ·················· | ·················· | ·················· | ·················· | ·················· |
| F | 9172 hCoV-19/South Korea/KCDC2652/2020\|EPI_ISL_515054\|2 | ·················· | ·················· | ·················· | ·················· | ·················· | ·················· |
| F | 9173 hCoV-19/Belgium/ULG-10248/2020\|EPI_ISL_515055\|2020 | ·················· | ·················· | ·················· | ·················· | ·················· | ·················· |
| F | 9174 hCoV-19/Belgium/ULG-10249/2020\|EPI_ISL_515056\|2020 | ·················· | ·················· | ·················· | ·················· | ·················· | ·················· |
| F | 9175 hCoV-19/Belgium/ULG-10250/2020\|EPI_ISL_515057\|2020 | ·················· | ·················· | ·················· | ·················· | ·················· | ·················· |
| F | 9176 hCoV-19/Belgium/ULG-10251/2020\|EPI_ISL_515058\|2020 | ·················· | ·················· | ·················· | ·················· | ·················· | ·················· |
| F | 9177 hCoV-19/Belgium/ULG-10252/2020\|EPI_ISL_515059\|2020 | ·················· | ·················· | ·················· | ·················· | ·················· | ·················· |
| F | 9178 hCoV-19/Belgium/ULG-10253/2020\|EPI_ISL_515060\|2020 | ·················· | ·················· | ·················· | ·················· | ·················· | ·················· |
| F | 9179 hCoV-19/Belgium/ULG-10254/2020\|EPI_ISL_515061\|2020 | ·················· | ·················· | ·················· | ·················· | ·················· | ·················· |
| F | 9180 hCoV-19/Belgium/ULG-10255/2020\|EPI_ISL_515062\|2020 | ·················· | ·················· | ·················· | ·················· | ·················· | ·················· |
| F | 9181 hCoV-19/Belgium/ULG-10256/2020\|EPI_ISL_515063\|2020 | ·················· | ·················· | ·················· | ·················· | ·················· | ·················· |
| F | 9182 hCoV-19/Belgium/ULG-10257/2020\|EPI_ISL_515064\|2020 | ·················· | ·················· | ·················· | ·················· | ·················· | ·················· |
| F | 9183 hCoV-19/Belgium/ULG-10258/2020\|EPI_ISL_515065\|2020 | ·················· | ·················· | ·················· | ·················· | ·················· | ·················· |
| F | 9184 hCoV-19/Belgium/ULG-10259/2020\|EPI_ISL_515066\|2020 | ·················· | ·················· | ·················· | ·················· | ·················· | ·················· |
| F | 9185 hCoV-19/Belgium/ULG-10260/2020\|EPI_ISL_515067\|2020 | ·················· | ·················· | ·················· | ·················· | ·················· | ·················· |
| F | 9186 hCoV-19/Belgium/ULG-10261/2020\|EPI_ISL_515068\|2020 | ·················· | ·················· | ·················· | ·················· | ·················· | ·················· |
| F | 9187 hCoV-19/Belgium/ULG-10262/2020\|EPI_ISL_515069\|2020 | ·················· | ·················· | ·················· | ·················· | ·················· | ·················· |
| F | 9188 hCoV-19/Belgium/ULG-10263/2020\|EPI_ISL_515070\|2020 | ·················· | ·················· | ·················· | ·················· | ·················· | ·················· |
| F | 9189 hCoV-19/Belgium/ULG-10264/2020\|EPI_ISL_515071\|2020 | ·················· | ·················· | ·················· | ·················· | ·················· | ·················· |
| F | 9190 hCoV-19/Belgium/ULG-10266/2020\|EPI_ISL_515073\|2020 | ·················· | ·················· | ·················· | ·················· | ·················· | ·················· |
| F | 9191 hCoV-19/Belgium/ULG-10267/2020\|EPI_ISL_515074\|2020 | ·················· | ·················· | ·················· | ·················· | ·················· | ·················· |
| F | 9192 hCoV-19/Belgium/ULG-10268/2020\|EPI_ISL_515075\|2020 | ·················· | ·················· | ·················· | ·················· | ·················· | ·················· |
| F | 9193 hCoV-19/Belgium/ULG-10269/2020\|EPI_ISL_515076\|2020 | ·················· | ·················· | ·················· | ·················· | ·················· | ·················· |
| F | 9194 hCoV-19/Belgium/ULG-10270/2020\|EPI_ISL_515077\|2020 | ·················· | ·················· | ·················· | ·················· | ·················· | ·················· |
| F | 9195 hCoV-19/Belgium/ULG-10271/2020\|EPI_ISL_515078\|2020 | ·················· | ·················· | ·················· | ·················· | ·················· | ·················· |
| F | 9196 hCoV-19/Belgium/ULG-10272/2020\|EPI_ISL_515079\|2020 | ·················· | ·················· | ·················· | ·················· | ·················· | ·················· |
| F | 9197 hCoV-19/Belgium/ULG-10273/2020\|EPI_ISL_515080\|2020 | ·················· | ·················· | ·················· | ·················· | ·················· | ·················· |
| F | 9198 hCoV-19/Belgium/ULG-10274/2020\|EPI_ISL_515081\|2020 | ·················· | ·················· | ·················· | ·················· | ·················· | ·················· |
| F | 9201 hCoV-19/Ghana/35012_S17/2020\|EPI_ISL_515090\|2020-0 | ·················· | ·················· | ·················· | ·················· | ·················· | ·················· |
| F | 9202 hCoV-19/Ghana/35563_S37/2020\|EPI_ISL_515096\|2020-0 | ·················· | ·················· | ·················· | ·················· | N················· | ·················· |
| F | 9204 hCoV-19/USA/MI-MDHHS-SC21624/2020\|EPI_ISL_516351\|2 | ·················· | ·················· | ·················· | ·················· | ·················· | ·················· |
| F | 9205 hCoV-19/USA/MI-MDHHS-SC21632/2020\|EPI_ISL_516359\|2 | ·················· | ·················· | ·················· | ·················· | ·················· | ·················· |
| F | 9206 hCoV-19/USA/MI-MDHHS-SC21633/2020\|EPI_ISL_516360\|2 | ·················· | ·················· | ·················· | ·················· | ·················· | ·················· |
| F | 9207 hCoV-19/USA/MI-MDHHS-SC21634/2020\|EPI_ISL_516361\|2 | ·················· | ·················· | ·················· | ·················· | ·················· | ·················· |
| F | 9208 hCoV-19/USA/MI-MDHHS-SC21635/2020\|EPI_ISL_516362\|2 | ·················· | ·················· | ·················· | ·················· | ·················· | ·················· |
| F | 9209 hCoV-19/USA/MI-MDHHS-SC21636/2020\|EPI_ISL_516363\|2 | ·················· | ·················· | ·················· | ·················· | ·················· | ·················· |
| F | 9210 hCoV-19/USA/MI-MDHHS-SC21638/2020\|EPI_ISL_516365\|2 | ·················· | ·················· | ·················· | ·················· | ·················· | ·················· |
| F | 9211 hCoV-19/USA/MI-MDHHS-SC21640/2020\|EPI_ISL_516367\|2 | ·················· | ·················· | ·················· | ·················· | ·················· | ·················· |
| F | 9212 hCoV-19/USA/MI-MDHHS-SC21644/2020\|EPI_ISL_516371\|2 | ·················· | ·················· | ·················· | ·················· | ·················· | ·················· |
| F | 9213 hCoV-19/USA/MI-MDHHS-SC21645/2020\|EPI_ISL_516372\|2 | ·················· | ·················· | ·················· | ·················· | ·················· | ·················· |
| F | 9214 hCoV-19/USA/MI-MDHHS-SC21646/2020\|EPI_ISL_516373\|2 | ·················· | ·················· | ·················· | ·················· | ·················· | ·················· |
| F | 9215 hCoV-19/USA/MI-MDHHS-SC21649/2020\|EPI_ISL_516376\|2 | ·················· | ·················· | ·················· | ·················· | ·················· | ·················· |
| F | 9216 hCoV-19/USA/MI-MDHHS-SC21653/2020\|EPI_ISL_516380\|2 | ·················· | ·················· | ·················· | ·················· | ·················· | ·················· |
| F | 9217 hCoV-19/South Korea/KCDC2099/2020\|EPI_ISL_498000\|2 | ·················· | ·················· | ·················· | ·················· | ·················· | ·················· |
| F | 9218 hCoV-19/South Korea/KCDC2100/2020\|EPI_ISL_498001\|2 | ·················· | ·················· | ·················· | ·················· | ·················· | ·················· |
| F | 9219 hCoV-19/South Korea/KCDC2101/2020\|EPI_ISL_498002\|2 | ·················· | ·················· | ·················· | ·················· | ·················· | ·················· |
| F | 9220 hCoV-19/South Korea/KCDC2102/2020\|EPI_ISL_498003\|2 | ·················· | ·················· | ·················· | ·················· | ·················· | ·················· |
| F | 9221 hCoV-19/South Korea/KCDC2103/2020\|EPI_ISL_498004\|2 | ·················· | ·················· | ·················· | ·················· | ·················· | ·················· |
| F | 9222 hCoV-19/South Korea/KCDC2104/2020\|EPI_ISL_498005\|2 | ·················· | ·················· | ·················· | ·················· | ·················· | ·················· |
| F | 9223 hCoV-19/South Korea/KCDC2105/2020\|EPI_ISL_498006\|2 | ·················· | ·················· | ·················· | ·················· | ·················· | ·················· |
| F | 9224 hCoV-19/South Korea/KCDC2106/2020\|EPI_ISL_498007\|2 | ·················· | ·················· | ·················· | ·················· | ·················· | ·················· |
| F | 9225 hCoV-19/South Korea/KCDC2107/2020\|EPI_ISL_498008\|2 | ·················· | ·················· | ·················· | ·················· | ·················· | ·················· |
| F | 9226 hCoV-19/South Korea/KCDC2108/2020\|EPI_ISL_498009\|2 | ·················· | ·················· | ·················· | ·················· | ·················· | ·················· |
| F | 9227 hCoV-19/South Korea/KCDC2109/2020\|EPI_ISL_498010\|2 | ·················· | ·················· | ·················· | ·················· | ·················· | ·················· |
| F | 9228 hCoV-19/South Korea/KCDC2110/2020\|EPI_ISL_498011\|2 | ·················· | ·················· | ·················· | ·················· | ·················· | ·················· |
| F | 9229 hCoV-19/South Korea/KCDC2111/2020\|EPI_ISL_498012\|2 | ·················· | ·················· | ·················· | ·················· | ·················· | ·················· |
| F | 9230 hCoV-19/South Korea/KCDC2112/2020\|EPI_ISL_498013\|2 | ·················· | ·················· | ·················· | ·················· | ·················· | ·················· |
| F | 9231 hCoV-19/South Korea/KCDC2113/2020\|EPI_ISL_498014\|2 | ·················· | ·················· | ·················· | ·················· | ·················· | ·················· |
| F | 9232 hCoV-19/South Korea/KCDC2114/2020\|EPI_ISL_498015\|2 | ·················· | ·················· | ·················· | ·················· | ·················· | ·················· |
| F | 9233 hCoV-19/South Korea/KCDC2115/2020\|EPI_ISL_498016\|2 | ·················· | ·················· | ·················· | ·················· | ·················· | ·················· |
| F | 9234 hCoV-19/South Korea/KCDC2119/2020\|EPI_ISL_498020\|2 | ·················· | ·················· | ·················· | ·················· | ·················· | ·················· |
| F | 9235 hCoV-19/South Korea/KCDC2120/2020\|EPI_ISL_498021\|2 | ·················· | ·················· | ·················· | ·················· | ·················· | ·················· |
| F | 9236 hCoV-19/South Korea/KCDC2121/2020\|EPI_ISL_498022\|2 | ·················· | ·················· | ·················· | ·················· | ·················· | ·················· |
| F | 9237 hCoV-19/South Korea/KCDC2122/2020\|EPI_ISL_498023\|2 | ·················· | ·················· | ·················· | ·················· | ·················· | ·················· |
| F | 9238 hCoV-19/South Korea/KCDC2123/2020\|EPI_ISL_498024\|2 | ·················· | ·················· | ·················· | ·················· | ·················· | ·················· |
| F | 9239 hCoV-19/South Korea/KCDC2124/2020\|EPI_ISL_498025\|2 | ·················· | ·················· | ·················· | ·················· | ·················· | ·················· |
| F | 9240 hCoV-19/South Korea/KCDC2125/2020\|EPI_ISL_498026\|2 | ·················· | ·················· | ·················· | ·················· | ·················· | ·················· |
| F | 9241 hCoV-19/South Korea/KCDC2137/2020\|EPI_ISL_498038\|2 | ·················· | ·················· | ·················· | ·················· | ·················· | ·················· |
| F | 9242 hCoV-19/South Korea/KCDC2138/2020\|EPI_ISL_498039\|2 | ·················· | ·················· | ·················· | ·················· | ·················· | ·················· |
| F | 9243 hCoV-19/South Korea/KCDC2139/2020\|EPI_ISL_498040\|2 | ·················· | ·················· | ·················· | ·················· | ·················· | ·················· |
| F | 9244 hCoV-19/South Korea/KCDC2140/2020\|EPI_ISL_498041\|2 | ·················· | ·················· | ·················· | ·················· | ·················· | ·················· |
| F | 9245 hCoV-19/South Korea/KCDC2141/2020\|EPI_ISL_498042\|2 | ·················· | ·················· | ·················· | ·················· | ·················· | ·················· |
| F | 9246 hCoV-19/South Korea/KCDC2142/2020\|EPI_ISL_498043\|2 | ·················· | ·················· | ·················· | ·················· | ·················· | ·················· |

| | | | | | | | | |
|---|---|---|---|---|---|---|---|---|
| F | 9247 hCoV-19/South Korea/KCDC2143/2020\|EPI_ISL_498044\|2 | ···················· | ···················· | ···················· | ···················· | ···················· | ···················· | |
| F | 9248 hCoV-19/South Korea/KCDC2144/2020\|EPI_ISL_498045\|2 | ···················· | ···················· | ···················· | ···················· | ···················· | ···················· | |
| F | 9249 hCoV-19/South Korea/KCDC2145/2020\|EPI_ISL_498046\|2 | ···················· | ···················· | ···················· | ···················· | ···················· | ···················· | |
| F | 9250 hCoV-19/South Korea/KCDC2146/2020\|EPI_ISL_498047\|2 | ···················· | ···················· | ···················· | ···················· | ···················· | ···················· | |
| F | 9251 hCoV-19/South Korea/KCDC2147/2020\|EPI_ISL_498048\|2 | ···················· | ···················· | ···················· | ···················· | ···················· | ···················· | |
| F | 9252 hCoV-19/South Korea/KCDC2148/2020\|EPI_ISL_498049\|2 | ···················· | ···················· | ···················· | ···················· | ···················· | ···················· | |
| F | 9253 hCoV-19/South Korea/KCDC2149/2020\|EPI_ISL_498050\|2 | ···················· | ···················· | ···················· | ···················· | ···················· | ···················· | |
| F | 9254 hCoV-19/South Korea/KCDC2150/2020\|EPI_ISL_498051\|2 | ···················· | ···················· | ···················· | ···················· | ···················· | ···················· | |
| F | 9255 hCoV-19/South Korea/KCDC2151/2020\|EPI_ISL_498052\|2 | ···················· | ···················· | ···················· | ···················· | ···················· | ···················· | |
| F | 9256 hCoV-19/South Korea/KCDC2152/2020\|EPI_ISL_498053\|2 | ···················· | ···················· | ···················· | ···················· | ···················· | ···················· | |
| F | 9257 hCoV-19/South Africa/KRISP-0436/2020\|EPI_ISL_49805 | ···················· | ···················· | ···················· | ···················· | ···················· | ···················· | |
| F | 9258 hCoV-19/England/PORT-2D221E/2020\|EPI_ISL_499303\|20 | ···················· | ····T··············· | ···················· | ···················· | ···················· | ···················· | |
| F | 9259 hCoV-19/England/PORT-2D222D/2020\|EPI_ISL_499304\|20 | ···················· | ···················· | ···················· | ···················· | ···················· | ···················· | |
| F | 9260 hCoV-19/England/PORT-2D22B4/2020\|EPI_ISL_499306\|20 | ···················· | ···················· | ···················· | ···················· | ···················· | ···················· | |
| F | 9261 hCoV-19/England/PORT-2D22C3/2020\|EPI_ISL_499307\|20 | ···················· | ···················· | ···················· | ···················· | ···················· | ···················· | |
| F | 9262 hCoV-19/England/PORT-2D22D2/2020\|EPI_ISL_499308\|20 | ···················· | ···················· | ···················· | ···················· | ···················· | ···················· | |
| F | 9263 hCoV-19/England/PORT-2D3AF7/2020\|EPI_ISL_499309\|20 | ···················· | ···················· | ···················· | ···················· | ···················· | ···················· | |
| F | 9264 hCoV-19/England/PORT-2D3B03/2020\|EPI_ISL_499310\|20 | ···················· | ···················· | ···················· | ···················· | ···················· | ···················· | |
| F | 9265 hCoV-19/England/PORT-2D3B21/2020\|EPI_ISL_499311\|20 | ···················· | ···················· | ···················· | ···················· | ···················· | ···················· | |
| F | 9266 hCoV-19/England/PORT-2D3B4F/2020\|EPI_ISL_499312\|20 | ···················· | ···················· | ···················· | ···················· | ···················· | ···················· | |
| F | 9267 hCoV-19/England/PORT-2D3BB8/2020\|EPI_ISL_499314\|20 | ···················· | ···················· | ···················· | ···················· | ···················· | ···················· | |
| F | 9268 hCoV-19/England/PORT-2D3C2E/2020\|EPI_ISL_499315\|20 | ···················· | ···················· | ···················· | ···················· | ···················· | ···················· | |
| F | 9269 hCoV-19/England/PORT-2D3C3D/2020\|EPI_ISL_499316\|20 | ···················· | ···················· | ···················· | ···················· | ···················· | ···················· | |
| F | 9270 hCoV-19/England/PORT-2D3C79/2020\|EPI_ISL_499318\|20 | ···················· | ···················· | ···················· | ···················· | ···················· | ···················· | |
| F | 9271 hCoV-19/England/PORT-2D3CF1/2020\|EPI_ISL_499322\|20 | ···················· | ···················· | ···················· | ···················· | ···················· | ···················· | |
| F | 9272 hCoV-19/England/PORT-2D3D1C/2020\|EPI_ISL_499323\|20 | ···················· | ···················· | ···················· | ···················· | ···················· | ···················· | |
| F | 9273 hCoV-19/England/PORT-2D3D2B/2020\|EPI_ISL_499324\|20 | ···················· | ···················· | ···················· | ···················· | ···················· | ···················· | |
| F | 9274 hCoV-19/England/PORT-2D3D3A/2020\|EPI_ISL_499325\|20 | ···················· | ···················· | ···················· | ···················· | ···················· | ···················· | |
| F | 9275 hCoV-19/England/PORT-2D3F07/2020\|EPI_ISL_499327\|20 | ···················· | ···················· | ···················· | ···················· | ···················· | ···················· | |
| F | 9276 hCoV-19/England/PORT-2D3F43/2020\|EPI_ISL_499328\|20 | ···················· | ···················· | ···················· | ···················· | ···················· | ···················· | |
| F | 9277 hCoV-19/England/PORT-2D3F52/2020\|EPI_ISL_499329\|20 | ···················· | ···················· | ···················· | ···················· | ···················· | ···················· | |
| F | 9278 hCoV-19/England/SHEF-C292D/2020\|EPI_ISL_499330\|202 | ···················· | ···················· | ···················· | ···················· | ···················· | ···················· | |
| F | 9279 hCoV-19/England/SHEF-C3FC0/2020\|EPI_ISL_499332\|202 | ···················· | ···················· | ···················· | ···················· | ···················· | ···················· | |
| F | 9280 hCoV-19/England/SHEF-C6473/2020\|EPI_ISL_499336\|202 | ···················· | ···················· | ···················· | ···················· | ···················· | ···················· | |
| F | 9281 hCoV-19/England/SHEF-CBFC8/2020\|EPI_ISL_499344\|202 | ···················· | ···················· | ···················· | ···················· | ···················· | ···················· | |
| F | 9282 hCoV-19/England/SHEF-CE9B8/2020\|EPI_ISL_499348\|202 | ···················· | ···················· | ···················· | ···················· | ···················· | ···················· | |
| F | 9283 hCoV-19/Scotland/CVR3956/2020\|EPI_ISL_499354\|2020- | ···················· | ···················· | ···················· | ···················· | ···················· | ···················· | |
| F | 9284 hCoV-19/Wales/PHWC-168E38/2020\|EPI_ISL_499355\|2020 | ···················· | ····A··············· | ···················· | ···················· | ···················· | ···················· | |
| F | 9285 hCoV-19/Wales/PHWC-168E47/2020\|EPI_ISL_499356\|2020 | ···················· | ···················· | ···················· | ···················· | ···················· | ···················· | |
| F | 9286 hCoV-19/Wales/PHWC-168E65/2020\|EPI_ISL_499357\|2020 | ···················· | ···················· | ···················· | ···················· | ···················· | ···················· | |
| F | 9287 hCoV-19/Wales/PHWC-168F35/2020\|EPI_ISL_499369\|2020 | ···················· | ···················· | ···················· | ···················· | ···················· | ···················· | |
| F | 9288 hCoV-19/Wales/PHWC-168F44/2020\|EPI_ISL_499370\|2020 | ···················· | ···················· | ···················· | ···················· | ···················· | ···················· | |
| F | 9289 hCoV-19/Wales/PHWC-168F62/2020\|EPI_ISL_499372\|2020 | ···················· | ···················· | ···················· | ···················· | ···················· | ···················· | |
| F | 9290 hCoV-19/Wales/PHWC-168F80/2020\|EPI_ISL_499374\|2020 | ···················· | ···················· | ···················· | ···················· | ···················· | ···················· | |
| F | 9291 hCoV-19/Wales/PHWC-168FAE/2020\|EPI_ISL_499375\|2020 | ···················· | ···················· | ···················· | ···················· | ···················· | ···················· | |
| F | 9292 hCoV-19/Wales/PHWC-168FBD/2020\|EPI_ISL_499376\|2020 | ···················· | ···················· | ···················· | ···················· | ···················· | ···················· | |
| F | 9293 hCoV-19/Wales/PHWC-168FCC/2020\|EPI_ISL_499377\|2020 | ···················· | ···················· | ···················· | ···················· | ···················· | ···················· | |
| F | 9294 hCoV-19/Wales/PHWC-168FDB/2020\|EPI_ISL_499378\|2020 | ···················· | ···················· | ···················· | ···················· | ···················· | ···················· | |
| F | 9295 hCoV-19/Wales/PHWC-168FEA/2020\|EPI_ISL_499379\|2020 | ···················· | ···················· | ···················· | ···················· | ···················· | ···················· | |
| F | 9296 hCoV-19/Wales/PHWC-168FF9/2020\|EPI_ISL_499380\|2020 | ···················· | ···················· | ···················· | ···················· | ···················· | ···················· | |
| F | 9297 hCoV-19/Wales/PHWC-169004/2020\|EPI_ISL_499381\|2020 | ···················· | ···················· | ···················· | ···················· | NNN················· | ···················· | 1 |
| F | 9298 hCoV-19/Wales/PHWC-169013/2020\|EPI_ISL_499382\|2020 | ···················· | ···················· | ···················· | ···················· | ···················· | ···················· | |
| F | 9299 hCoV-19/Wales/PHWC-169031/2020\|EPI_ISL_499384\|2020 | ···················· | ···················· | ···················· | ···················· | ···················· | ···················· | |
| F | 9300 hCoV-19/Wales/PHWC-169040/2020\|EPI_ISL_499385\|2020 | ···················· | ···················· | ···················· | ···················· | ···················· | ···················· | |
| F | 9302 hCoV-19/Wales/PHWC-16906E/2020\|EPI_ISL_499387\|2020 | ···················· | ···················· | ···················· | ···················· | ···················· | ···················· | |
| F | 9303 hCoV-19/Wales/PHWC-16907D/2020\|EPI_ISL_499388\|2020 | ···················· | ···················· | ···················· | ···················· | NNN················· | ···················· | 1 |
| F | 9305 hCoV-19/Australia/NSW2017/2020\|EPI_ISL_500601\|2020 | ···················· | ···················· | ···················· | ···················· | ···················· | ···················· | |
| F | 9306 hCoV-19/Australia/NSW2018/2020\|EPI_ISL_500602\|2020 | ···················· | ···················· | ···················· | ···················· | ···················· | ···················· | |
| F | 9307 hCoV-19/Australia/NSW2020/2020\|EPI_ISL_500603\|2020 | ···················· | ···················· | ···················· | ···················· | ···················· | ···················· | |
| F | 9308 hCoV-19/Australia/NSW2021/2020\|EPI_ISL_500604\|2020 | ···················· | ···················· | ···················· | ···················· | ···················· | ···················· | |
| F | 9309 hCoV-19/Australia/NSW2026/2020\|EPI_ISL_500605\|2020 | ···················· | ···················· | ···················· | ···················· | ···················· | ···················· | |
| F | 9310 hCoV-19/Australia/NSW2027/2020\|EPI_ISL_500606\|2020 | ···················· | ···················· | ···················· | ···················· | ···················· | ···················· | |
| F | 9311 hCoV-19/Australia/NSW2032/2020\|EPI_ISL_500607\|2020 | ···················· | ·············A······ | ···················· | ···················· | ···················· | ···················· | |
| F | 9312 hCoV-19/Australia/NSW2033/2020\|EPI_ISL_500608\|2020 | ···················· | ·············A······ | ···················· | ···················· | ···················· | ···················· | |
| F | 9313 hCoV-19/Australia/NSW2037/2020\|EPI_ISL_500609\|2020 | ···················· | ·············A······ | ···················· | ···················· | ···················· | ···················· | |
| F | 9314 hCoV-19/Australia/NSW2038/2020\|EPI_ISL_500610\|2020 | ···················· | ·············A······ | ···················· | ···················· | ···················· | ···················· | |
| F | 9315 hCoV-19/Australia/NSW2039/2020\|EPI_ISL_500611\|2020 | ···················· | ·············A······ | ···················· | ···················· | ···················· | ···················· | |
| F | 9316 hCoV-19/Australia/NSW2041/2020\|EPI_ISL_500612\|2020 | ···················· | ···················· | ···················· | ···················· | ···················· | ···················· | |
| F | 9317 hCoV-19/Australia/NSW2042/2020\|EPI_ISL_500613\|2020 | ···················· | ···················· | ···················· | ···················· | ···················· | ···················· | |
| F | 9318 hCoV-19/Australia/NSW2043/2020\|EPI_ISL_500614\|2020 | ···················· | ···················· | ···················· | ···················· | ···················· | ···················· | |
| F | 9319 hCoV-19/Australia/NSW2044/2020\|EPI_ISL_500615\|2020 | ···················· | ···················· | ···················· | ···················· | ···················· | ···················· | |
| F | 9320 hCoV-19/Australia/NSW2045/2020\|EPI_ISL_500616\|2020 | ···················· | ···················· | ···················· | ···················· | ···················· | ···················· | |
| F | 9321 hCoV-19/Australia/NSW2046/2020\|EPI_ISL_500617\|2020 | ···················· | ·············N······ | ···················· | ···················· | ···················· | ···················· | |
| F | 9322 hCoV-19/Australia/NSW2047/2020\|EPI_ISL_500618\|2020 | ···················· | ···················· | ···················· | ···················· | ···················· | ···················· | |
| F | 9323 hCoV-19/Australia/NSW2049/2020\|EPI_ISL_500619\|2020 | ···················· | ···················· | ···················· | ···················· | ···················· | ···················· | |
| F | 9324 hCoV-19/Australia/NSW2050/2020\|EPI_ISL_500620\|2020 | ···················· | ···················· | ···················· | ···················· | ···················· | ···················· | |
| F | 9325 hCoV-19/Australia/NSW2115/2020\|EPI_ISL_500621\|2020 | ···················· | ···················· | ···················· | ···················· | ···················· | ···················· | |
| F | 9326 hCoV-19/Australia/NSW2116/2020\|EPI_ISL_500622\|2020 | ···················· | ···················· | ···················· | ···················· | ···················· | ···················· | |
| F | 9327 hCoV-19/Australia/NSW2128/2020\|EPI_ISL_500623\|2020 | ···················· | ···················· | ···················· | ···················· | ···················· | ···················· | |
| F | 9328 hCoV-19/Australia/NSW2131/2020\|EPI_ISL_500624\|2020 | ···················· | ···················· | ···················· | ···················· | ···················· | ···················· | |

| | | | | | | | | |
|---|---|---|---|---|---|---|---|---|
| F | 9329 | hCoV-19/Australia/NSW2132/2020\|EPI_ISL_500625\|2020 | ···················· | ·A·················· | ···················· | ···················· | ···················· | ···················· |
| F | 9330 | hCoV-19/Australia/NSW2137/2020\|EPI_ISL_500626\|2020 | ···················· | ···················· | ···················· | ···················· | ···················· | ···················· |
| F | 9331 | hCoV-19/Australia/NSW2139/2020\|EPI_ISL_500627\|2020 | ···················· | ···················· | ···················· | ···················· | ···················· | ···················· |
| F | 9332 | hCoV-19/Australia/NSW2147/2020\|EPI_ISL_500628\|2020 | ···················· | ···················· | ···················· | ···················· | ···················· | ···················· |
| F | 9333 | hCoV-19/Australia/NSW2148/2020\|EPI_ISL_500629\|2020 | ···················· | ···················· | ···················· | ···················· | ···················· | ···················· |
| F | 9334 | hCoV-19/Australia/NSW2149/2020\|EPI_ISL_500630\|2020 | ···················· | ···················· | ···················· | ···················· | ···················· | ···················· |
| F | 9335 | hCoV-19/Australia/NSW2151/2020\|EPI_ISL_500631\|2020 | ···················· | ···················· | ···················· | ···················· | ···················· | ···················· |
| F | 9336 | hCoV-19/Australia/NSW2154/2020\|EPI_ISL_500632\|2020 | ···················· | ···················· | ···················· | ···················· | ···················· | ···················· |
| F | 9337 | hCoV-19/Australia/NSW2155/2020\|EPI_ISL_500633\|2020 | ···················· | ···················· | ···················· | ···················· | ···················· | ···················· |
| F | 9338 | hCoV-19/Australia/NSW2156/2020\|EPI_ISL_500634\|2020 | ···················· | ···················· | ···················· | ···················· | ···················· | ···················· |
| F | 9339 | hCoV-19/Australia/NSW2157/2020\|EPI_ISL_500635\|2020 | ···················· | ···················· | ···················· | ···················· | ···················· | ···················· |
| F | 9340 | hCoV-19/Australia/NSW2158/2020\|EPI_ISL_500636\|2020 | ···················· | ···················· | ···················· | ···················· | ···················· | ···················· |
| F | 9341 | hCoV-19/Australia/NSW2159/2020\|EPI_ISL_500637\|2020 | ···················· | ···················· | ···················· | ···················· | ···················· | ···················· |
| F | 9342 | hCoV-19/Australia/NSW2164/2020\|EPI_ISL_500638\|2020 | ···················· | ···················· | ···················· | ···················· | ···················· | ···················· |
| F | 9343 | hCoV-19/Australia/NSW2168/2020\|EPI_ISL_500639\|2020 | ···················· | ···················· | ···················· | ···················· | ···················· | ···················· |
| F | 9344 | hCoV-19/Australia/NSW2169/2020\|EPI_ISL_500640\|2020 | ···················· | ···················· | ···················· | ···················· | ···················· | ···················· |
| F | 9345 | hCoV-19/Australia/NSW2170/2020\|EPI_ISL_500641\|2020 | ···················· | ···················· | ···················· | ···················· | ···················· | ···················· |
| F | 9346 | hCoV-19/Australia/NSW2171/2020\|EPI_ISL_500642\|2020 | ···················· | ···················· | ···················· | ···················· | ···················· | ···················· |
| F | 9347 | hCoV-19/Australia/NSW2172/2020\|EPI_ISL_500643\|2020 | ···················· | ···················· | ···················· | ···················· | ···················· | ···················· |
| F | 9348 | hCoV-19/Australia/NSW2173/2020\|EPI_ISL_500644\|2020 | ···················· | ···················· | ···················· | ···················· | ···················· | ···················· |
| F | 9349 | hCoV-19/Australia/NSW2174/2020\|EPI_ISL_500645\|2020 | ···················· | ···················· | ···················· | ···················· | ···················· | ···················· |
| F | 9350 | hCoV-19/Australia/NSW2175/2020\|EPI_ISL_500646\|2020 | ···················· | ···················· | ···················· | ···················· | ···················· | ···················· |
| F | 9351 | hCoV-19/Australia/NSW2176/2020\|EPI_ISL_500647\|2020 | ···················· | ···················· | ···················· | ···················· | ···················· | ···················· |
| F | 9352 | hCoV-19/Australia/NSW2181/2020\|EPI_ISL_500648\|2020 | ···················· | ···················· | ···················· | ···················· | ···················· | ···················· |
| F | 9353 | hCoV-19/Australia/NSW2185/2020\|EPI_ISL_500649\|2020 | ···················· | ···················· | ···················· | ···················· | ···················· | ···················· |
| F | 9354 | hCoV-19/Australia/NSW2187/2020\|EPI_ISL_500650\|2020 | ···················· | ···················· | ···················· | ···················· | ···················· | ···················· |
| F | 9355 | hCoV-19/Australia/NSW2188/2020\|EPI_ISL_500651\|2020 | ···················· | ···················· | ···················· | ···················· | ···················· | ···················· |
| F | 9356 | hCoV-19/Australia/NSW2189/2020\|EPI_ISL_500652\|2020 | ···················· | ···················· | ···················· | ···················· | ···················· | ···················· |
| F | 9357 | hCoV-19/Australia/NSW2191/2020\|EPI_ISL_500653\|2020 | ···················· | ···················· | ···················· | ···················· | ···················· | ···················· |
| F | 9358 | hCoV-19/Australia/NSW2192/2020\|EPI_ISL_500654\|2020 | ···················· | ···················· | ···················· | ···················· | ···················· | ···················· |
| F | 9359 | hCoV-19/Australia/NSW2194/2020\|EPI_ISL_500655\|2020 | ···················· | ···················· | ···················· | ···················· | ···················· | ···················· |
| F | 9360 | hCoV-19/Australia/NSW2195/2020\|EPI_ISL_500656\|2020 | ···················· | ···················· | ···················· | ···················· | ···················· | ···················· |
| F | 9361 | hCoV-19/Australia/NSW2196/2020\|EPI_ISL_500657\|2020 | ···················· | ···················· | ···················· | ···················· | ···················· | ···················· |
| F | 9362 | hCoV-19/Australia/NSW2197/2020\|EPI_ISL_500658\|2020 | ···················· | ···················· | ···················· | ···················· | ···················· | ···················· |
| F | 9363 | hCoV-19/Australia/NSW2198/2020\|EPI_ISL_500659\|2020 | ···················· | ···················· | ···················· | ···················· | ···················· | ···················· |
| F | 9364 | hCoV-19/Australia/NSW2199/2020\|EPI_ISL_500660\|2020 | ···················· | ·A·················· | ···················· | ···················· | ···················· | ···················· |
| F | 9365 | hCoV-19/Australia/NSW2201/2020\|EPI_ISL_500661\|2020 | ···················· | ···················· | ···················· | ···················· | ···················· | ···················· |
| F | 9366 | hCoV-19/Australia/NSW2202/2020\|EPI_ISL_500662\|2020 | ···················· | ···················· | ···················· | ···················· | ···················· | ···················· |
| F | 9367 | hCoV-19/Australia/NSW2203/2020\|EPI_ISL_500663\|2020 | ···················· | ···················· | ···················· | ···················· | ···················· | ···················· |
| F | 9368 | hCoV-19/Australia/NSW2206/2020\|EPI_ISL_500664\|2020 | ···················· | ···················· | ···················· | ···················· | ···················· | ···················· |
| F | 9369 | hCoV-19/Australia/NSW2207/2020\|EPI_ISL_500665\|2020 | ···················· | ···················· | ···················· | ···················· | ···················· | ···················· |
| F | 9370 | hCoV-19/Australia/NSW2208/2020\|EPI_ISL_500666\|2020 | ···················· | ···················· | ···················· | ···················· | ···················· | ···················· |
| F | 9371 | hCoV-19/Australia/NSW2231/2020\|EPI_ISL_500678\|2020 | ···················· | ···················· | ···················· | ···················· | ···················· | ···················· |
| F | 9372 | hCoV-19/Australia/NSW2237/2020\|EPI_ISL_500679\|2020 | ···················· | ···················· | ···················· | ···················· | ···················· | ···················· |
| F | 9373 | hCoV-19/Australia/NSW2239/2020\|EPI_ISL_500681\|2020 | ···················· | ···················· | ···················· | ···················· | ···················· | ···················· |
| F | 9374 | hCoV-19/Australia/NSW2242/2020\|EPI_ISL_500682\|2020 | ···················· | ···················· | ···················· | ···················· | ···················· | ···················· |
| F | 9375 | hCoV-19/Australia/NSW2243/2020\|EPI_ISL_500683\|2020 | ···················· | ···················· | ···················· | ···················· | ···················· | ···················· |
| F | 9376 | hCoV-19/Australia/NSW2244/2020\|EPI_ISL_500684\|2020 | ···················· | ···················· | ···················· | ···················· | ···················· | ···················· |
| F | 9377 | hCoV-19/Australia/NSW2245/2020\|EPI_ISL_500685\|2020 | ···················· | ···················· | ···················· | ···················· | ···················· | ···················· |
| F | 9378 | hCoV-19/Australia/NSW2247/2020\|EPI_ISL_500686\|2020 | ···················· | ···················· | ···················· | ···················· | ···················· | ···················· |
| F | 9379 | hCoV-19/Australia/NSW2248/2020\|EPI_ISL_500687\|2020 | ···················· | ···················· | ···················· | ···················· | ···················· | ···················· |
| F | 9380 | hCoV-19/Australia/NSW2249/2020\|EPI_ISL_500688\|2020 | ···················· | ···················· | ···················· | ···················· | ···················· | ···················· |
| F | 9381 | hCoV-19/Latvia/087/2020\|EPI_ISL_501936\|2020-07-09 | ···················· | ···················· | ···················· | ···················· | ···················· | ···················· |
| F | 9382 | hCoV-19/France/ARA-02052/2020\|EPI_ISL_509010\|2020- | ···················· | ···················· | ···················· | ···················· | ···················· | ···················· |
| F | 9383 | hCoV-19/France/ARA-17037/2020\|EPI_ISL_509011\|2020- | ···················· | ···················· | ···················· | ···················· | ···················· | ···················· |
| F | 9384 | hCoV-19/France/ARA-17181/2020\|EPI_ISL_509012\|2020- | ···················· | ···················· | ···················· | ···················· | ···················· | ···················· |
| F | 9385 | hCoV-19/France/ARA-17410/2020\|EPI_ISL_509013\|2020- | ···················· | ···················· | ···················· | ···················· | ···················· | ···················· |
| F | 9386 | hCoV-19/France/ARA-17573/2020\|EPI_ISL_509014\|2020- | ···················· | ···················· | ···················· | ···················· | ···················· | ···················· |
| F | 9387 | hCoV-19/France/ARA-18066/2020\|EPI_ISL_509015\|2020- | ···················· | ···················· | ···················· | ···················· | ···················· | ···················· |
| F | 9388 | hCoV-19/France/ARA-18148/2020\|EPI_ISL_509016\|2020- | ···················· | ···················· | ···················· | ···················· | ···················· | ···················· |
| F | 9389 | hCoV-19/Poland/A_348r/2020\|EPI_ISL_509017\|2020-06 | ···················· | ···················· | ···················· | ···················· | ···················· | ···················· |
| F | 9390 | hCoV-19/Poland/B_348r/2020\|EPI_ISL_509018\|2020-06 | ···················· | ···················· | ···················· | ···················· | ···················· | ···················· |
| F | 9391 | hCoV-19/USA/OR-OHSU-0165/2020\|EPI_ISL_509066\|2020- | ···················· | ···················· | ···················· | ···················· | ···················· | ···················· |
| F | 9392 | hCoV-19/USA/OR-OHSU-0166/2020\|EPI_ISL_509067\|2020- | ···················· | ···················· | ···················· | ···················· | ···················· | ···················· |
| F | 9393 | hCoV-19/USA/OR-OHSU-0190/2020\|EPI_ISL_509069\|2020- | ···················· | ···················· | ···················· | ···················· | ···················· | ···················· |
| F | 9394 | hCoV-19/USA/OR-OHSU-0245/2020\|EPI_ISL_509080\|2020- | ···················· | ···················· | ···················· | ···················· | ···················· | ···················· |
| F | 9395 | hCoV-19/USA/OR-OHSU-0247/2020\|EPI_ISL_509081\|2020- | ···················· | ···················· | ···················· | ···················· | ···················· | ···················· |
| F | 9396 | hCoV-19/USA/OR-OHSU-0256/2020\|EPI_ISL_509082\|2020- | ···················· | ···················· | ···················· | ···················· | ···················· | ···················· |
| F | 9397 | hCoV-19/USA/OR-OHSU-0258/2020\|EPI_ISL_509083\|2020- | ···················· | ···················· | ···················· | ···················· | ···················· | ···················· |
| F | 9398 | hCoV-19/USA/OR-OHSU-0260/2020\|EPI_ISL_509084\|2020- | ···················· | ···················· | ···················· | ···················· | ···················· | ···················· |
| F | 9399 | hCoV-19/USA/OR-OHSU-0265/2020\|EPI_ISL_509085\|2020- | ···················· | ···················· | ···················· | ···················· | ···················· | ···················· |
| F | 9400 | hCoV-19/USA/OR-OHSU-0267/2020\|EPI_ISL_509086\|2020- | ···················· | ···················· | ···················· | ···················· | ···················· | ···················· |
| F | 9401 | hCoV-19/USA/OR-OHSU-0269/2020\|EPI_ISL_509087\|2020- | ···················· | ···················· | ···················· | ···················· | ···················· | ···················· |
| F | 9402 | hCoV-19/USA/OR-OHSU-0273/2020\|EPI_ISL_509088\|2020- | ···················· | ···················· | ···················· | ···················· | ···················· | ···················· |
| F | 9403 | hCoV-19/USA/OR-OHSU-0276/2020\|EPI_ISL_509089\|2020- | ···················· | ···················· | ···················· | ···················· | ···················· | ···················· |
| F | 9404 | hCoV-19/USA/OR-OHSU-0279/2020\|EPI_ISL_509090\|2020- | ···················· | ···················· | ···················· | ···················· | ···················· | ···················· |
| F | 9405 | hCoV-19/USA/OR-OHSU-0281/2020\|EPI_ISL_509091\|2020- | ···················· | ···················· | ···················· | ···················· | ···················· | ···················· |
| F | 9406 | hCoV-19/USA/OR-OHSU-0292/2020\|EPI_ISL_509092\|2020- | ···················· | ···················· | ···················· | ···················· | ···················· | ···················· |
| F | 9407 | hCoV-19/USA/OR-OHSU-0293/2020\|EPI_ISL_509093\|2020- | ···················· | ···················· | ···················· | ···················· | ···················· | ···················· |
| F | 9408 | hCoV-19/USA/OR-OHSU-0294/2020\|EPI_ISL_509094\|2020- | ···················· | ···················· | ···················· | ···················· | ···················· | ···················· |

| | | | | | | | | |
|---|---|---|---|---|---|---|---|---|
| F | 9409 | hCoV-19/USA/OR-OHSU-0301/2020\|EPI_ISL_509095\|2020- | ···················· | ···················· | ···················· | ···················· | ···················· | ···················· |
| F | 9410 | hCoV-19/USA/OR-OHSU-0303/2020\|EPI_ISL_509097\|2020- | ···················· | ···················· | ···················· | ···················· | ···················· | ···················· |
| F | 9411 | hCoV-19/USA/OR-OHSU-0308/2020\|EPI_ISL_509099\|2020- | ···················· | ···················· | ···················· | ···················· | ···················· | ···················· |
| F | 9415 | hCoV-19/Portugal/PT1342/2020\|EPI_ISL_511301\|2020-0 | ···················· | ···················· | ···················· | ···················· | ···················· | ···················· |
| F | 9416 | hCoV-19/Portugal/PT1343/2020\|EPI_ISL_511302\|2020-0 | ···················· | ···················· | ···················· | ···················· | ···················· | ···················· |
| F | 9417 | hCoV-19/Portugal/PT1348/2020\|EPI_ISL_511307\|2020-0 | ···················· | ···················· | ···················· | ···················· | ···················· | ···················· |
| F | 9418 | hCoV-19/Portugal/PT1351/2020\|EPI_ISL_511310\|2020-0 | ···················· | ···················· | ···················· | ···················· | ···················· | ···················· |
| F | 9419 | hCoV-19/Portugal/PT1352/2020\|EPI_ISL_511311\|2020-0 | ···················· | ···················· | ···················· | ···················· | ···················· | ···················· |
| F | 9420 | hCoV-19/Portugal/PT1353/2020\|EPI_ISL_511312\|2020-0 | ···················· | ···················· | ···················· | ···················· | ···················· | ···················· |
| F | 9421 | hCoV-19/Portugal/PT1354/2020\|EPI_ISL_511313\|2020-0 | ···················· | ···················· | ···················· | ···················· | ···················· | ···················· |
| F | 9422 | hCoV-19/Portugal/PT1355/2020\|EPI_ISL_511314\|2020-0 | ···················· | ···················· | ···················· | ···················· | ···················· | ···················· |
| F | 9423 | hCoV-19/Portugal/PT1357/2020\|EPI_ISL_511316\|2020-0 | ···················· | ···················· | ···················· | ···················· | ···················· | ···················· |
| F | 9424 | hCoV-19/Portugal/PT1365/2020\|EPI_ISL_511324\|2020-0 | ···················· | ···················· | ···················· | ···················· | ···················· | ···················· |
| F | 9425 | hCoV-19/Ukraine/203100320/2020\|EPI_ISL_512600\|2020 | ···················· | ···················· | ···················· | ···················· | ···················· | ···················· |
| F | 9426 | hCoV-19/Ukraine/203100323/2020\|EPI_ISL_512603\|2020 | ···················· | ···················· | ···················· | ···················· | ···················· | ···················· |
| F | 9427 | hCoV-19/Ukraine/203100324/2020\|EPI_ISL_512604\|2020 | ···················· | ···················· | ···················· | ···················· | ···················· | ···················· |
| F | 9428 | hCoV-19/Ukraine/203100325/2020\|EPI_ISL_512605\|2020 | ···················· | ···················· | ···················· | ···················· | ···················· | ···················· |
| F | 9429 | hCoV-19/Ukraine/203100327/2020\|EPI_ISL_512606\|2020 | ···················· | ···················· | ···················· | ···················· | ···················· | ···················· |
| F | 9430 | hCoV-19/Ukraine/203100328/2020\|EPI_ISL_512607\|2020 | ···················· | ···················· | ···················· | ···················· | ···················· | ···················· |
| F | 9431 | hCoV-19/Ukraine/203100329/2020\|EPI_ISL_512608\|2020 | ···················· | ···················· | ···················· | ···················· | ···················· | ···················· |
| F | 9432 | hCoV-19/Ukraine/203100330/2020\|EPI_ISL_512609\|2020 | ···················· | ···················· | ···················· | ···················· | ···················· | ···················· |
| F | 9433 | hCoV-19/Ukraine/203100331/2020\|EPI_ISL_512610\|2020 | ···················· | ···················· | ···················· | ···················· | ···················· | ···················· |
| F | 9434 | hCoV-19/Ukraine/203100332/2020\|EPI_ISL_512611\|2020 | ···················· | ···················· | ···················· | ···················· | ···················· | ···················· |
| F | 9435 | hCoV-19/Ukraine/203100333/2020\|EPI_ISL_512612\|2020 | ···················· | ···················· | ···················· | ···················· | ···················· | ···················· |
| F | 9436 | hCoV-19/Ukraine/203100335/2020\|EPI_ISL_512614\|2020 | ···················· | ···················· | ···················· | ···················· | ···················· | ···················· |
| F | 9437 | hCoV-19/Ukraine/203100336/2020\|EPI_ISL_512615\|2020 | ···················· | ···················· | ···················· | ···················· | ···················· | ···················· |
| F | 9438 | hCoV-19/Ukraine/203100337/2020\|EPI_ISL_512616\|2020 | ···················· | ···················· | ···················· | ···················· | ···················· | ···················· |
| F | 9439 | hCoV-19/Ukraine/203100338/2020\|EPI_ISL_512617\|2020 | ···················· | ···················· | ···················· | ···················· | ···················· | ···················· |
| F | 9440 | hCoV-19/Ukraine/203100339/2020\|EPI_ISL_512618\|2020 | ···················· | ···················· | ···················· | ···················· | ···················· | ···················· |
| F | 9441 | hCoV-19/Ukraine/203100341/2020\|EPI_ISL_512620\|2020 | ···················· | ···················· | ···················· | ···················· | ···················· | ···················· |
| F | 9442 | hCoV-19/Ukraine/203100342/2020\|EPI_ISL_512621\|2020 | ···················· | ···················· | ···················· | ···················· | ···················· | ···················· |
| F | 9443 | hCoV-19/Ukraine/203100343/2020\|EPI_ISL_512622\|2020 | ···················· | ···················· | ···················· | ···················· | ···················· | ···················· |
| F | 9444 | hCoV-19/Ukraine/203100344/2020\|EPI_ISL_512623\|2020 | ···················· | ···················· | ···················· | ···················· | ···················· | ···················· |
| F | 9445 | hCoV-19/Ukraine/203100345/2020\|EPI_ISL_512624\|2020 | ···················· | ···················· | ···················· | ···················· | ···················· | ···················· |
| F | 9446 | hCoV-19/Ukraine/203100346/2020\|EPI_ISL_512625\|2020 | ···················· | ···················· | ···················· | ···················· | ···················· | ···················· |
| F | 9447 | hCoV-19/Ukraine/203100347/2020\|EPI_ISL_512626\|2020 | ···················· | ···················· | ···················· | ···················· | ···················· | ···················· |
| F | 9448 | hCoV-19/Ukraine/203100349/2020\|EPI_ISL_512628\|2020 | ···················· | ···················· | ···················· | ···················· | ···················· | ···················· |
| F | 9449 | hCoV-19/Ukraine/203100350/2020\|EPI_ISL_512629\|2020 | ···················· | ···················· | ···················· | ···················· | ···················· | ···················· |
| F | 9450 | hCoV-19/Ukraine/203100352/2020\|EPI_ISL_512631\|2020 | ···················· | ···················· | ···················· | ···················· | ···················· | ···················· |
| F | 9451 | hCoV-19/Ukraine/203100353/2020\|EPI_ISL_512632\|2020 | ···················· | ···················· | ···················· | ···················· | ···················· | ···················· |
| F | 9452 | hCoV-19/Ukraine/203100355/2020\|EPI_ISL_512634\|2020 | ···················· | ···················· | ···················· | ···················· | ···················· | ···················· |
| F | 9453 | hCoV-19/Ukraine/203100357/2020\|EPI_ISL_512636\|2020 | ···················· | ···················· | ···················· | ···················· | ···················· | ···················· |
| F | 9454 | hCoV-19/Ukraine/203100359/2020\|EPI_ISL_512638\|2020 | ···················· | ···················· | ···················· | ···················· | ···················· | ···················· |
| F | 9455 | hCoV-19/Ukraine/203100361/2020\|EPI_ISL_512640\|2020 | ···················· | ···················· | ···················· | ···················· | ···················· | ···················· |
| F | 9456 | hCoV-19/Ukraine/203100363/2020\|EPI_ISL_512641\|2020 | ···················· | ···················· | ···················· | ···················· | ···················· | ···················· |
| F | 9457 | hCoV-19/Ukraine/203100364/2020\|EPI_ISL_512642\|2020 | ···················· | ···················· | ···················· | ···················· | ···················· | ···················· |
| F | 9458 | hCoV-19/Ukraine/203100365/2020\|EPI_ISL_512643\|2020 | ···················· | ···················· | ···················· | ···················· | ···················· | ···················· |
| F | 9459 | hCoV-19/Latvia/090/2020\|EPI_ISL_512645\|2020-07-22 | ···················· | ···················· | ···················· | ···················· | ···················· | ···················· |
| F | 9460 | hCoV-19/Latvia/091/2020\|EPI_ISL_512646\|2020-07-07 | ···················· | ···················· | ···················· | ···················· | ···················· | ···················· |
| F | 9461 | hCoV-19/Latvia/092/2020\|EPI_ISL_512647\|2020-06-26 | ···················· | ···················· | ···················· | ···················· | ···················· | ···················· |
| F | 9462 | hCoV-19/Latvia/093/2020\|EPI_ISL_512648\|2020-07-14 | ···················· | ···················· | ···················· | ···················· | ···················· | ···················· |
| F | 9463 | hCoV-19/Latvia/094/2020\|EPI_ISL_512649\|2020-07-21 | ···················· | ···················· | ···················· | ···················· | ···················· | ···················· |
| F | 9464 | hCoV-19/Latvia/095/2020\|EPI_ISL_512650\|2020-07-21 | ···················· | ···················· | ···················· | ···················· | ···················· | ···················· |
| F | 9465 | hCoV-19/Latvia/096/2020\|EPI_ISL_512651\|2020-07-23 | ···················· | ·A·················· | ···················· | ···················· | ···················· | ···················· |
| F | 9466 | hCoV-19/Latvia/097/2020\|EPI_ISL_512652\|2020-07-25 | ···················· | ···················· | ···················· | ···················· | ···················· | ···················· |
| F | 9467 | hCoV-19/Costa Rica/INC-0047/2020\|EPI_ISL_512653\|20 | ···················· | ···················· | ···················· | ···················· | ···················· | ···················· |
| F | 9468 | hCoV-19/Costa Rica/INC-0048/2020\|EPI_ISL_512654\|20 | ···················· | ···················· | ···················· | ···················· | ···················· | ···················· |
| F | 9469 | hCoV-19/Costa Rica/INC-0049/2020\|EPI_ISL_512655\|20 | ···················· | ···················· | ···················· | ···················· | ···················· | ···················· |
| F | 9470 | hCoV-19/Costa Rica/INC-0050/2020\|EPI_ISL_512656\|20 | ···················· | ···················· | ···················· | ···················· | ···················· | ···················· |
| F | 9471 | hCoV-19/Australia/SAP481/2020\|EPI_ISL_513911\|2020- | ···················· | ···················· | ···················· | ···················· | ···················· | ···················· |
| F | 9472 | hCoV-19/Pakistan/KPK-KUST-SJTU/2020\|EPI_ISL_513925 | ···················· | ···················· | ···················· | ···················· | ···················· | ···················· |
| F | 9473 | hCoV-19/England/PORT-2D1D2D/2020\|EPI_ISL_499272\|20 | ···················· | ···················· | ···················· | ···················· | ···················· | ···················· |
| F | 9474 | hCoV-19/England/PORT-2D1DA5/2020\|EPI_ISL_499278\|20 | ···················· | ···················· | ···················· | ···················· | ···················· | ···················· |
| F | 9475 | hCoV-19/England/PORT-2D1DE1/2020\|EPI_ISL_499281\|20 | ···················· | ···················· | ···················· | ···················· | ···················· | ···················· |
| F | 9476 | hCoV-19/England/PORT-2D1E1B/2020\|EPI_ISL_499282\|20 | ···················· | ···················· | ···················· | ···················· | ···················· | ···················· |
| F | 9477 | hCoV-19/England/PORT-2D1E2A/2020\|EPI_ISL_499283\|20 | ···················· | ···················· | ···················· | ···················· | ···················· | ···················· |
| F | 9478 | hCoV-19/England/PORT-2D1E48/2020\|EPI_ISL_499285\|20 | ···················· | ···················· | ···················· | ···················· | ···················· | ···················· |
| F | 9479 | hCoV-19/England/PORT-2D1E84/2020\|EPI_ISL_499288\|20 | ···················· | ···················· | ···················· | ···················· | ···················· | ···················· |
| F | 9480 | hCoV-19/England/PORT-2D1E93/2020\|EPI_ISL_499289\|20 | ···················· | ···················· | ···················· | ···················· | ···················· | ···················· |
| F | 9481 | hCoV-19/England/PORT-2D1EFD/2020\|EPI_ISL_499294\|20 | ···················· | ···················· | ···················· | ···················· | ···················· | ···················· |
| F | 9482 | hCoV-19/England/PORT-2D1F09/2020\|EPI_ISL_499295\|20 | ···················· | ····T··············· | ···················· | ···················· | ···················· | ···················· |
| F | 9483 | hCoV-19/England/PORT-2D1F18/2020\|EPI_ISL_499296\|20 | ···················· | ···················· | ···················· | ···················· | ···················· | ···················· |
| F | 9484 | hCoV-19/England/PORT-2D1F27/2020\|EPI_ISL_499297\|20 | ···················· | ···················· | ···················· | ···················· | ···················· | ···················· |
| F | 9485 | hCoV-19/England/PORT-2D1F36/2020\|EPI_ISL_499298\|20 | ···················· | ···················· | ···················· | ···················· | ···················· | ···················· |
| F | 9486 | hCoV-19/England/PORT-2D1F72/2020\|EPI_ISL_499300\|20 | ···················· | ···················· | ···················· | ···················· | ···················· | ···················· |
| F | 9487 | hCoV-19/England/PORT-2D1F90/2020\|EPI_ISL_499302\|20 | ···················· | ···················· | ···················· | ···················· | ···················· | ···················· |
| F | 9488 | hCoV-19/Australia/NSW2250/2020\|EPI_ISL_500689\|2020 | ···················· | ···················· | ···················· | ···················· | ···················· | ···················· |
| F | 9489 | hCoV-19/Australia/NSW2251/2020\|EPI_ISL_500690\|2020 | ···················· | ···················· | ···················· | ···················· | ···················· | ···················· |
| F | 9490 | hCoV-19/Australia/NSW2252/2020\|EPI_ISL_500691\|2020 | ···················· | ···················· | ···················· | ···················· | ···················· | ···················· |
| F | 9491 | hCoV-19/Australia/NSW2253/2020\|EPI_ISL_500692\|2020 | ···················· | ···················· | ···················· | ···················· | ···················· | ···················· |

| | | | | | | | | |
|---|---|---|---|---|---|---|---|---|
| F | 9492 | hCoV-19/Australia/NSW2256/2020\|EPI_ISL_500693\|2020 | .................... | .................... | .................... | .................... | .................... | .................... |
| F | 9493 | hCoV-19/Australia/NSW2257/2020\|EPI_ISL_500694\|2020 | .................... | .................... | .................... | .................... | .................... | .................... |
| F | 9494 | hCoV-19/Australia/NSW2258/2020\|EPI_ISL_500695\|2020 | .................... | .................... | .................... | .................... | .................... | .................... |
| F | 9495 | hCoV-19/Australia/NSW2259/2020\|EPI_ISL_500696\|2020 | .................... | .................... | .................... | .................... | .................... | .................... |
| F | 9496 | hCoV-19/Australia/NSW2261/2020\|EPI_ISL_500697\|2020 | .................... | .................... | .................... | .................... | .................... | .................... |
| F | 9497 | hCoV-19/Australia/NSW2262/2020\|EPI_ISL_500698\|2020 | .................... | .................... | .................... | .................... | .................... | .................... |
| F | 9498 | hCoV-19/Australia/NSW2263/2020\|EPI_ISL_500699\|2020 | .................... | .................... | .................... | .................... | .................... | .................... |
| F | 9499 | hCoV-19/Australia/NSW2265/2020\|EPI_ISL_500700\|2020 | .................... | .................... | .................... | .................... | .................... | .................... |
| F | 9500 | hCoV-19/Australia/NSW2266/2020\|EPI_ISL_500701\|2020 | .................... | .................... | .................... | .................... | .................... | .................... |
| F | 9501 | hCoV-19/Australia/NSW2283/2020\|EPI_ISL_500702\|2020 | .................... | .................... | .................... | .................... | .................... | .................... |
| F | 9502 | hCoV-19/Australia/NSW2292/2020\|EPI_ISL_500703\|2020 | .................... | .................... | .................... | .................... | .................... | .................... |
| F | 9503 | hCoV-19/Australia/NSW2306/2020\|EPI_ISL_500704\|2020 | .................... | .................... | .................... | .................... | .................... | .................... |
| F | 9504 | hCoV-19/Australia/NSW2310/2020\|EPI_ISL_500705\|2020 | .................... | .................... | .................... | .................... | .................... | .................... |
| F | 9505 | hCoV-19/Australia/NSW2314/2020\|EPI_ISL_500706\|2020 | .................... | .................... | .................... | .................... | .................... | .................... |
| F | 9506 | hCoV-19/England/203020503/2020\|EPI_ISL_500709\|2020 | .................... | .................... | .................... | .................... | .................... | .................... |
| F | 9507 | hCoV-19/England/203020504/2020\|EPI_ISL_500710\|2020 | .................... | .................... | .................... | .................... | .................... | .................... |
| F | 9508 | hCoV-19/England/203020505/2020\|EPI_ISL_500711\|2020 | .................... | .................... | .................... | .................... | .................... | .................... |
| F | 9509 | hCoV-19/England/203020514/2020\|EPI_ISL_500715\|2020 | .................... | .................... | .................... | .................... | .................... | .................... |
| F | 9510 | hCoV-19/Australia/NSW2608/2020\|EPI_ISL_500717\|2020 | .................... | .................... | .................... | .................... | .................... | .................... |
| F | 9511 | hCoV-19/Poland/E_348r/2020\|EPI_ISL_509020\|2020-06 | .................... | .................... | .................... | .................... | .................... | .................... |
| F | 9512 | hCoV-19/Poland/F_348r/2020\|EPI_ISL_509021\|2020-06 | .................... | .................... | .................... | .................... | .................... | .................... |
| F | 9513 | hCoV-19/Poland/G_348r/2020\|EPI_ISL_509022\|2020-06 | .................... | .................... | .................... | .................... | .................... | .................... |
| F | 9514 | hCoV-19/Poland/H_348r/2020\|EPI_ISL_509023\|2020-06 | .................... | .................... | .................... | .................... | .................... | .................... |
| F | 9515 | hCoV-19/Poland/I_348r/2020\|EPI_ISL_509024\|2020-06 | .................... | .................... | .................... | .................... | .................... | .................... |
| F | 9516 | hCoV-19/Poland/J_348r/2020\|EPI_ISL_509026\|2020-06 | .................... | .................... | .................... | .................... | .................... | .................... |
| F | 9517 | hCoV-19/Poland/K_348r/2020\|EPI_ISL_509028\|2020-06 | .................... | .................... | .................... | .................... | .................... | .................... |
| F | 9518 | hCoV-19/Poland/L_348r/2020\|EPI_ISL_509029\|2020-06 | .................... | .................... | .................... | .................... | .................... | .................... |
| F | 9519 | hCoV-19/Poland/M_348r/2020\|EPI_ISL_509031\|2020-06 | .................... | .................... | .................... | .................... | .................... | .................... |
| F | 9520 | hCoV-19/Poland/O_348r/2020\|EPI_ISL_509033\|2020-06 | .................... | .................... | .................... | .................... | .................... | .................... |
| F | 9521 | hCoV-19/Poland/P_348r/2020\|EPI_ISL_509035\|2020-06 | .................... | .................... | .................... | .................... | .................... | .................... |
| F | 9522 | hCoV-19/Poland/S_348r/2020\|EPI_ISL_509037\|2020-06 | .................... | .................... | .................... | .................... | .................... | .................... |
| F | 9523 | hCoV-19/Poland/W_348r/2020\|EPI_ISL_509038\|2020-06 | .................... | .................... | .................... | .................... | .................... | .................... |
| F | 9524 | hCoV-19/Poland/X_348r/2020\|EPI_ISL_509051\|2020-06 | .................... | .................... | .................... | .................... | .................... | .................... |
| F | 9525 | hCoV-19/USA/OR-OHSU-0066/2020\|EPI_ISL_509059\|2020- | .................... | .................... | .................... | .................... | .................... | .................... |
| F | 9526 | hCoV-19/USA/OR-OHSU-0091/2020\|EPI_ISL_509060\|2020- | .................... | .................... | .................... | .................... | .................... | .................... |
| F | 9527 | hCoV-19/USA/OR-OHSU-0106/2020\|EPI_ISL_509061\|2020- | .................... | .................... | .................... | .................... | .................... | .................... |
| F | 9528 | hCoV-19/USA/OR-OHSU-0110/2020\|EPI_ISL_509062\|2020- | .................... | .................... | .................... | .................... | .................... | .................... |
| F | 9529 | hCoV-19/USA/OR-OHSU-0161/2020\|EPI_ISL_509064\|2020- | .................... | .................... | .................... | .................... | .................... | .................... |
| F | 9530 | hCoV-19/USA/OR-OHSU-0162/2020\|EPI_ISL_509065\|2020- | .................... | .................... | .................... | .................... | .................... | .................... |
| F | 9531 | hCoV-19/USA/OR-OHSU-0193/2020\|EPI_ISL_509070\|2020- | .................... | .................... | .................... | .................... | .................... | .................... |
| F | 9532 | hCoV-19/USA/OR-OHSU-0200/2020\|EPI_ISL_509071\|2020- | .................... | .................... | .................... | .................... | .................... | .................... |
| F | 9533 | hCoV-19/USA/OR-OHSU-0215/2020\|EPI_ISL_509072\|2020- | .................... | .................... | .................... | .................... | .................... | .................... |
| F | 9534 | hCoV-19/USA/OR-OHSU-0221/2020\|EPI_ISL_509073\|2020- | .................... | .................... | .................... | .................... | .................... | .................... |
| F | 9535 | hCoV-19/USA/OR-OHSU-0222/2020\|EPI_ISL_509074\|2020- | .................... | .................... | .................... | .................... | .................... | .................... |
| F | 9536 | hCoV-19/USA/OR-OHSU-0232/2020\|EPI_ISL_509075\|2020- | .................... | .................... | .................... | .................... | .................... | .................... |
| F | 9537 | hCoV-19/USA/OR-OHSU-0233/2020\|EPI_ISL_509076\|2020- | .................... | .................... | .................... | .................... | .................... | .................... |
| F | 9538 | hCoV-19/USA/OR-OHSU-0244/2020\|EPI_ISL_509079\|2020- | .................... | .................... | .................... | .................... | .................... | .................... |
| F | 9542 | hCoV-19/Spain/AN-IBV-004121/2020\|EPI_ISL_510058\|20 | .................... | .................... | .................... | .................... | .................... | .................... |
| F | 9548 | hCoV-19/Portugal/PT1378/2020\|EPI_ISL_511337\|2020-0 | .................... | .................... | .................... | .................... | .................... | .................... |
| F | 9549 | hCoV-19/Portugal/PT1379/2020\|EPI_ISL_511338\|2020-0 | .................... | .................... | .................... | .................... | .................... | .................... |
| F | 9550 | hCoV-19/Portugal/PT1380/2020\|EPI_ISL_511339\|2020-0 | .................... | .................... | .................... | .................... | .................... | .................... |
| F | 9551 | hCoV-19/Portugal/PT1382/2020\|EPI_ISL_511341\|2020-0 | .................... | .................... | .................... | .................... | .................... | .................... |
| F | 9552 | hCoV-19/Portugal/PT1384/2020\|EPI_ISL_511343\|2020-0 | .................... | .................... | .................... | .................... | .................... | .................... |
| F | 9553 | hCoV-19/Portugal/PT1386/2020\|EPI_ISL_511345\|2020-0 | .................... | .................... | .................... | .................... | .................... | .................... |
| F | 9554 | hCoV-19/Portugal/PT1387/2020\|EPI_ISL_511346\|2020-0 | .................... | .................... | .................... | .................... | .................... | .................... |
| F | 9555 | hCoV-19/Portugal/PT1388/2020\|EPI_ISL_511347\|2020-0 | .................... | .................... | .................... | .................... | .................... | .................... |
| F | 9556 | hCoV-19/Portugal/PT1389/2020\|EPI_ISL_511348\|2020-0 | .................... | .................... | .................... | .................... | .................... | .................... |
| F | 9557 | hCoV-19/Portugal/PT1391/2020\|EPI_ISL_511350\|2020-0 | .................... | .................... | .................... | .................... | .................... | .................... |
| F | 9558 | hCoV-19/Portugal/PT1392/2020\|EPI_ISL_511351\|2020-0 | .................... | .................... | .................... | .................... | .................... | .................... |
| F | 9559 | hCoV-19/Portugal/PT1393/2020\|EPI_ISL_511352\|2020-0 | .................... | .................... | .................... | .................... | .................... | .................... |
| F | 9560 | hCoV-19/Portugal/PT1394/2020\|EPI_ISL_511353\|2020-0 | .................... | .................... | .................... | .................... | .................... | .................... |
| F | 9561 | hCoV-19/Portugal/PT1396/2020\|EPI_ISL_511355\|2020-0 | .................... | .................... | .................... | .................... | .................... | .................... |
| F | 9562 | hCoV-19/Portugal/PT1398/2020\|EPI_ISL_511357\|2020-0 | .................... | .................... | .................... | .................... | .................... | .................... |
| F | 9563 | hCoV-19/Portugal/PT1399/2020\|EPI_ISL_511358\|2020-0 | .................... | .................... | .................... | .................... | .................... | .................... |
| F | 9564 | hCoV-19/Portugal/PT1400/2020\|EPI_ISL_511359\|2020-0 | .................... | .................... | .................... | .................... | .................... | .................... |
| F | 9565 | hCoV-19/Portugal/PT1401/2020\|EPI_ISL_511360\|2020-0 | .................... | .................... | .................... | .................... | .................... | .................... |
| F | 9566 | hCoV-19/Portugal/PT1402/2020\|EPI_ISL_511361\|2020-0 | .................... | .................... | .................... | .................... | .................... | .................... |
| F | 9567 | hCoV-19/Portugal/PT1403/2020\|EPI_ISL_511362\|2020-0 | .................... | .................... | .................... | .................... | .................... | .................... |
| F | 9568 | hCoV-19/Portugal/PT1404/2020\|EPI_ISL_511363\|2020-0 | .................... | .................... | .................... | .................... | .................... | .................... |
| F | 9569 | hCoV-19/Portugal/PT1405/2020\|EPI_ISL_511364\|2020-0 | .................... | .................... | .................... | .................... | .................... | .................... |
| F | 9570 | hCoV-19/Portugal/PT1407/2020\|EPI_ISL_511366\|2020-0 | .................... | .................... | .................... | .................... | .................... | .................... |
| F | 9571 | hCoV-19/Portugal/PT1408/2020\|EPI_ISL_511367\|2020-0 | .................... | .................... | .................... | .................... | .................... | .................... |
| F | 9572 | hCoV-19/Portugal/PT1409/2020\|EPI_ISL_511368\|2020-0 | .................... | .................... | .................... | .................... | .................... | .................... |
| F | 9573 | hCoV-19/Portugal/PT1410/2020\|EPI_ISL_511369\|2020-0 | .................... | .................... | .................... | .................... | .................... | .................... |
| F | 9574 | hCoV-19/Portugal/PT1411/2020\|EPI_ISL_511370\|2020-0 | .................... | .................... | .................... | .................... | .................... | .................... |
| F | 9575 | hCoV-19/Portugal/PT1412/2020\|EPI_ISL_511371\|2020-0 | .................... | .................... | .................... | .................... | .................... | .................... |
| F | 9576 | hCoV-19/Portugal/PT1413/2020\|EPI_ISL_511372\|2020-0 | .................... | .................... | .................... | .................... | .................... | .................... |
| F | 9577 | hCoV-19/Portugal/PT1414/2020\|EPI_ISL_511373\|2020-0 | .................... | .................... | .................... | .................... | .................... | .................... |
| F | 9578 | hCoV-19/Portugal/PT1415/2020\|EPI_ISL_511374\|2020-0 | .................... | .................... | .................... | .................... | .................... | .................... |
| F | 9579 | hCoV-19/Portugal/PT1416/2020\|EPI_ISL_511375\|2020-0 | .................... | .................... | .................... | .................... | .................... | .................... |

| | | | | | | | | |
|---|---|---|---|---|---|---|---|---|
| F | 9580 | hCoV-19/Portugal/PT1417/2020\|EPI_ISL_511376\|2020-0 | .................... | .................... | .................... | .................... | .................... | .................... |
| F | 9581 | hCoV-19/Portugal/PT1418/2020\|EPI_ISL_511377\|2020-0 | .................... | .................... | .................... | .................... | .................... | .................... |
| F | 9582 | hCoV-19/Portugal/PT1419/2020\|EPI_ISL_511378\|2020-0 | .................... | .................... | .................... | .................... | .................... | .................... |
| F | 9583 | hCoV-19/Portugal/PT1420/2020\|EPI_ISL_511379\|2020-0 | .................... | .................... | .................... | .................... | .................... | .................... |
| F | 9584 | hCoV-19/Portugal/PT1422/2020\|EPI_ISL_511381\|2020-0 | .................... | .................... | .................... | .................... | .................... | .................... |
| F | 9585 | hCoV-19/Portugal/PT1423/2020\|EPI_ISL_511382\|2020-0 | .................... | .................... | .................... | .................... | .................... | .................... |
| F | 9586 | hCoV-19/Portugal/PT1424/2020\|EPI_ISL_511383\|2020-0 | .................... | .................... | .................... | .................... | .................... | .................... |
| F | 9587 | hCoV-19/Portugal/PT1425/2020\|EPI_ISL_511384\|2020-0 | .................... | .................... | .................... | .................... | .................... | .................... |
| F | 9588 | hCoV-19/Portugal/PT1426/2020\|EPI_ISL_511385\|2020-0 | .................... | .................... | .................... | .................... | .................... | .................... |
| F | 9589 | hCoV-19/Portugal/PT1427/2020\|EPI_ISL_511386\|2020-0 | .................... | .................... | .................... | .................... | .................... | .................... |
| F | 9590 | hCoV-19/Portugal/PT1428/2020\|EPI_ISL_511387\|2020-0 | .................... | .................... | .................... | .................... | .................... | .................... |
| F | 9591 | hCoV-19/Portugal/PT1429/2020\|EPI_ISL_511388\|2020-0 | .................... | .................... | .................... | .................... | .................... | .................... |
| F | 9592 | hCoV-19/Portugal/PT1430/2020\|EPI_ISL_511389\|2020-0 | .................... | .................... | .................... | .................... | .................... | .................... |
| F | 9593 | hCoV-19/Portugal/PT1431/2020\|EPI_ISL_511390\|2020-0 | .................... | .................... | .................... | .................... | .................... | .................... |
| F | 9594 | hCoV-19/Portugal/PT1432/2020\|EPI_ISL_511391\|2020-0 | .................... | .................... | .................... | .................... | .................... | .................... |
| F | 9595 | hCoV-19/Portugal/PT1433/2020\|EPI_ISL_511392\|2020-0 | .................... | .................... | .................... | .................... | .................... | .................... |
| F | 9596 | hCoV-19/Portugal/PT1434/2020\|EPI_ISL_511393\|2020-0 | .................... | .................... | .................... | .................... | .................... | .................... |
| F | 9597 | hCoV-19/Portugal/PT1435/2020\|EPI_ISL_511394\|2020-0 | .................... | .................... | .................... | .................... | .................... | .................... |
| F | 9598 | hCoV-19/Portugal/PT1436/2020\|EPI_ISL_511395\|2020-0 | .................... | .................... | .................... | .................... | .................... | .................... |
| F | 9599 | hCoV-19/Portugal/PT1437/2020\|EPI_ISL_511396\|2020-0 | .................... | .................... | .................... | .................... | .................... | .................... |
| F | 9600 | hCoV-19/Portugal/PT1438/2020\|EPI_ISL_511397\|2020-0 | .................... | .................... | .................... | .................... | .................... | .................... |
| F | 9601 | hCoV-19/Portugal/PT1439/2020\|EPI_ISL_511398\|2020-0 | .................... | .................... | .................... | .................... | .................... | .................... |
| F | 9602 | hCoV-19/Portugal/PT1440/2020\|EPI_ISL_511399\|2020-0 | .................... | .................... | .................... | .................... | .................... | .................... |
| F | 9603 | hCoV-19/Costa Rica/INC-0062/2020\|EPI_ISL_512668\|20 | .................... | .................... | .................... | .................... | .................... | .................... |
| F | 9604 | hCoV-19/Costa Rica/INC-0063/2020\|EPI_ISL_512669\|20 | .................... | .................... | .................... | .................... | .................... | .................... |
| F | 9605 | hCoV-19/Costa Rica/INC-0064/2020\|EPI_ISL_512670\|20 | .................... | .................... | .................... | .................... | .................... | .................... |
| F | 9606 | hCoV-19/Costa Rica/INC-0065/2020\|EPI_ISL_512671\|20 | .................... | .................... | .................... | .................... | .................... | .................... |
| F | 9607 | hCoV-19/Costa Rica/INC-0067/2020\|EPI_ISL_512672\|20 | .................... | .................... | .................... | .................... | .................... | .................... |
| F | 9608 | hCoV-19/Costa Rica/INC-0068/2020\|EPI_ISL_512673\|20 | .................... | .................... | .................... | .................... | .................... | .................... |
| F | 9609 | hCoV-19/Costa Rica/INC-0069/2020\|EPI_ISL_512674\|20 | .................... | .................... | .................... | .................... | .................... | .................... |
| F | 9610 | hCoV-19/Costa Rica/INC-0070/2020\|EPI_ISL_512675\|20 | .................... | .................... | .................... | .................... | .................... | .................... |
| F | 9612 | hCoV-19/USA/MI-MDHHS-SC21540/2020\|EPI_ISL_516267\|2 | .................... | .................... | .................... | .................... | .................... | .................... |
| F | 9613 | hCoV-19/USA/MI-MDHHS-SC21541/2020\|EPI_ISL_516268\|2 | .................... | .................... | .................... | .................... | .................... | .................... |
| F | 9614 | hCoV-19/USA/MI-MDHHS-SC21542/2020\|EPI_ISL_516269\|2 | .................... | .................... | .................... | .................... | .................... | .................... |
| F | 9615 | hCoV-19/USA/MI-MDHHS-SC21543/2020\|EPI_ISL_516270\|2 | .................... | .................... | .................... | .................... | .................... | .................... |
| F | 9616 | hCoV-19/USA/MI-MDHHS-SC21545/2020\|EPI_ISL_516272\|2 | .................... | .................... | .................... | .................... | .................... | .................... |
| F | 9617 | hCoV-19/USA/MI-MDHHS-SC21546/2020\|EPI_ISL_516273\|2 | .................... | .................... | .................... | .................... | .................... | .................... |
| F | 9618 | hCoV-19/USA/MI-MDHHS-SC21547/2020\|EPI_ISL_516274\|2 | .................... | .................... | .................... | .................... | .................... | .................... |
| F | 9619 | hCoV-19/USA/MI-MDHHS-SC21548/2020\|EPI_ISL_516275\|2 | .................... | .................... | .................... | .................... | .................... | .................... |
| F | 9620 | hCoV-19/USA/MI-MDHHS-SC21550/2020\|EPI_ISL_516277\|2 | .................... | .................... | .................... | .................... | .................... | .................... |
| F | 9621 | hCoV-19/USA/MI-MDHHS-SC21551/2020\|EPI_ISL_516278\|2 | .................... | .................... | .................... | .................... | .................... | .................... |
| F | 9622 | hCoV-19/USA/MI-MDHHS-SC21554/2020\|EPI_ISL_516281\|2 | .................... | .................... | .................... | .................... | .................... | .................... |
| F | 9623 | hCoV-19/USA/MI-MDHHS-SC21555/2020\|EPI_ISL_516282\|2 | .................... | .................... | .................... | .................... | .................... | .................... |
| F | 9624 | hCoV-19/USA/MI-MDHHS-SC21556/2020\|EPI_ISL_516283\|2 | .................... | .................... | .................... | .................... | .................... | .................... |
| F | 9625 | hCoV-19/USA/MI-MDHHS-SC21558/2020\|EPI_ISL_516285\|2 | .................... | .................... | .................... | .................... | .................... | .................... |
| F | 9626 | hCoV-19/USA/MI-MDHHS-SC21560/2020\|EPI_ISL_516287\|2 | .................... | .................... | .................... | .................... | .................... | .................... |
| F | 9627 | hCoV-19/USA/MI-MDHHS-SC21567/2020\|EPI_ISL_516294\|2 | .................... | .................... | .................... | .................... | .................... | .................... |
| F | 9628 | hCoV-19/USA/MI-MDHHS-SC21568/2020\|EPI_ISL_516295\|2 | .................... | .................... | .................... | .................... | .................... | .................... |
| F | 9629 | hCoV-19/USA/MI-MDHHS-SC21569/2020\|EPI_ISL_516296\|2 | .................... | .................... | .................... | .................... | .................... | .................... |
| F | 9630 | hCoV-19/USA/MI-MDHHS-SC21570/2020\|EPI_ISL_516297\|2 | .................... | .................... | .................... | .................... | .................... | .................... |
| G | 1 | hCoV-19/Israel/CVL-n19140/2020\|EPI_ISL_516900\|2020 | .................... | .................... | .................... | .................... | .................... | .................... |
| G | 2 | hCoV-19/Israel/CVL1060/2020\|EPI_ISL_516901\|2020-06 | .................... | .................... | .................... | .................... | .................... | .................... |
| G | 3 | hCoV-19/Israel/CVL1130/2020\|EPI_ISL_516902\|2020-06 | .................... | .................... | .................... | .................... | .................... | .................... |
| G | 4 | hCoV-19/Australia/VIC4712/2020\|EPI_ISL_519100\|2020 | .................... | .................... | .................... | .................... | .................... | .................... |
| G | 5 | hCoV-19/Australia/VIC4713/2020\|EPI_ISL_519101\|2020 | .................... | .................... | .................... | .................... | .................... | .................... |
| G | 6 | hCoV-19/Australia/VIC4714/2020\|EPI_ISL_519102\|2020 | .................... | .................... | .................... | .................... | .................... | .................... |
| G | 7 | hCoV-19/Australia/VIC4715/2020\|EPI_ISL_519103\|2020 | .................... | .................... | .................... | .................... | .................... | .................... |
| G | 8 | hCoV-19/Australia/VIC4716/2020\|EPI_ISL_519104\|2020 | .................... | .................... | .................... | .................... | .................... | .................... |
| G | 9 | hCoV-19/Australia/VIC4717/2020\|EPI_ISL_519105\|2020 | .................... | .................... | .................... | .................... | .................... | .................... |
| G | 10 | hCoV-19/Australia/VIC4718/2020\|EPI_ISL_519106\|2020 | .................... | .................... | .................... | .................... | .................... | .................... |
| G | 11 | hCoV-19/Australia/VIC4719/2020\|EPI_ISL_519107\|2020 | .................... | .................... | .................... | .................... | .................... | .................... |
| G | 12 | hCoV-19/Australia/VIC4720/2020\|EPI_ISL_519108\|2020 | .................... | .................... | .................... | .................... | .................... | .................... |
| G | 13 | hCoV-19/Australia/VIC4721/2020\|EPI_ISL_519109\|2020 | .................... | .................... | .................... | .................... | .................... | .................... |
| G | 14 | hCoV-19/Australia/VIC4722/2020\|EPI_ISL_519110\|2020 | .................... | .................... | .................... | .................... | .................... | .................... |
| G | 15 | hCoV-19/Australia/VIC4723/2020\|EPI_ISL_519111\|2020 | .................... | .................... | .................... | .................... | .................... | .................... |
| G | 16 | hCoV-19/Australia/VIC4725/2020\|EPI_ISL_519113\|2020 | .................... | .................... | .................... | .................... | .................... | .................... |
| G | 17 | hCoV-19/Australia/VIC4726/2020\|EPI_ISL_519114\|2020 | .................... | .................... | .................... | .................... | .................... | .................... |
| G | 18 | hCoV-19/Australia/VIC4727/2020\|EPI_ISL_519115\|2020 | .................... | .................... | .................... | .................... | .................... | .................... |
| G | 19 | hCoV-19/Australia/VIC4728/2020\|EPI_ISL_519116\|2020 | .................... | .................... | .................... | .................... | .................... | .................... |
| G | 20 | hCoV-19/Australia/VIC4729/2020\|EPI_ISL_519117\|2020 | .................... | .................... | .................... | .................... | .................... | .................... |
| G | 21 | hCoV-19/Australia/VIC4730/2020\|EPI_ISL_519118\|2020 | .................... | .................... | .................... | .................... | .................... | .................... |
| G | 22 | hCoV-19/Australia/VIC4731/2020\|EPI_ISL_519119\|2020 | .................... | .................... | .................... | .................... | .................... | .................... |
| G | 23 | hCoV-19/Australia/VIC4732/2020\|EPI_ISL_519120\|2020 | .................... | .................... | .................... | .................... | .................... | .................... |
| G | 24 | hCoV-19/Australia/VIC4733/2020\|EPI_ISL_519121\|2020 | .................... | .................... | .................... | .................... | .................... | .................... |
| G | 25 | hCoV-19/Australia/VIC4734/2020\|EPI_ISL_519122\|2020 | .................... | .................... | .................... | .................... | .................... | .................... |
| G | 26 | hCoV-19/Australia/VIC4735/2020\|EPI_ISL_519123\|2020 | .................... | .................... | .................... | .................... | .................... | .................... |
| G | 27 | hCoV-19/Australia/VIC4736/2020\|EPI_ISL_519124\|2020 | .................... | .................... | .................... | .................... | .................... | .................... |
| G | 28 | hCoV-19/Australia/VIC4737/2020\|EPI_ISL_519125\|2020 | .................... | .................... | .................... | .................... | .................... | .................... |
| G | 29 | hCoV-19/Australia/VIC4738/2020\|EPI_ISL_519126\|2020 | .................... | .................... | .................... | .................... | .................... | .................... |
| G | 30 | hCoV-19/Australia/VIC4739/2020\|EPI_ISL_519127\|2020 | .................... | .................... | .................... | .................... | .................... | .................... |

G 31 hCoV-19/Australia/VIC4740/2020|EPI_ISL_519128|2020 ···················· ···················· ····················· ···················· ···················· ····················
G 32 hCoV-19/Australia/VIC4742/2020|EPI_ISL_519130|2020 ···················· ···················· ····················· ···················· ···················· ····················
G 33 hCoV-19/Australia/VIC4743/2020|EPI_ISL_519131|2020 ···················· ···················· ····················· ···················· ···················· ····················
G 34 hCoV-19/Australia/VIC4744/2020|EPI_ISL_519132|2020 ···················· ···················· ····················· ···················· ···················· ····················
G 35 hCoV-19/Australia/VIC4745/2020|EPI_ISL_519133|2020 ···················· ···················· ····················· ···················· ···················· ····················
G 36 hCoV-19/Australia/VIC4746/2020|EPI_ISL_519134|2020 ···················· ···················· ····················· ···················· ···················· ····················
G 37 hCoV-19/Australia/VIC4747/2020|EPI_ISL_519135|2020 ···················· ···················· ····················· ···················· ···················· ····················
G 38 hCoV-19/Australia/VIC4748/2020|EPI_ISL_519136|2020 ···················· ···················· ····················· ···················· ···················· ····················
G 39 hCoV-19/Australia/VIC4749/2020|EPI_ISL_519137|2020 ···················· ···················· ····················· ···················· ···················· ····················
G 40 hCoV-19/Australia/VIC4750/2020|EPI_ISL_519138|2020 ···················· ···················· ····················· ···················· ···················· ····················
G 41 hCoV-19/Australia/VIC4751/2020|EPI_ISL_519139|2020 ···················· ···················· ····················· ···················· ···················· ····················
G 42 hCoV-19/Australia/VIC4752/2020|EPI_ISL_519140|2020 ···················· ···················· ····················· ···················· ···················· ····················
G 43 hCoV-19/Australia/VIC4753/2020|EPI_ISL_519141|2020 ···················· ···················· ····················· ···················· ···················· ····················
G 44 hCoV-19/Australia/VIC4754/2020|EPI_ISL_519142|2020 ···················· ···················· ····················· ···················· ···················· ····················
G 45 hCoV-19/Australia/VIC4755/2020|EPI_ISL_519143|2020 ···················· ···················· ····················· ···················· ···················· ····················
G 46 hCoV-19/Australia/VIC4756/2020|EPI_ISL_519144|2020 ···················· ···················· ····················· ···················· ···················· ····················
G 47 hCoV-19/Australia/VIC4757/2020|EPI_ISL_519145|2020 ···················· ···················· ····················· ···················· ···················· ····················
G 48 hCoV-19/Australia/VIC4758/2020|EPI_ISL_519146|2020 ···················· ···················· ····················· ···················· ···················· ····················
G 49 hCoV-19/Australia/VIC4759/2020|EPI_ISL_519147|2020 ···················· ···················· ····················· ···················· ···················· ····················
G 50 hCoV-19/Australia/VIC4760/2020|EPI_ISL_519148|2020 ···················· ···················· ····················· ···················· ···················· ····················
G 51 hCoV-19/Australia/VIC4761/2020|EPI_ISL_519149|2020 ···················· ···················· ····················· ···················· ···················· ····················
G 52 hCoV-19/Australia/VIC4762/2020|EPI_ISL_519150|2020 ···················· ···················· ····················· ···················· ···················· ····················
G 53 hCoV-19/Australia/VIC4763/2020|EPI_ISL_519151|2020 ···················· ···················· ····················· ···················· ···················· ····················
G 54 hCoV-19/Australia/VIC4764/2020|EPI_ISL_519152|2020 ···················· ···················· ····················· ···················· ···················· ····················
G 55 hCoV-19/Australia/VIC4765/2020|EPI_ISL_519153|2020 ···················· ···················· ····················· ···················· ···················· ····················
G 56 hCoV-19/Australia/VIC4766/2020|EPI_ISL_519154|2020 ···················· ···················· ····················· ···················· ···················· ····················
G 57 hCoV-19/Australia/VIC4767/2020|EPI_ISL_519155|2020 ···················· ···················· ····················· ···················· ···················· ····················
G 58 hCoV-19/Australia/VIC4768/2020|EPI_ISL_519156|2020 ···················· ···················· ····················· ···················· ···················· ····················
G 59 hCoV-19/Australia/VIC4769/2020|EPI_ISL_519157|2020 ···················· ···················· ····················· ···················· ···················· ····················
G 60 hCoV-19/Australia/VIC4772/2020|EPI_ISL_519158|2020 ···················· ···················· ····················· ···················· ···················· ····················
G 61 hCoV-19/Australia/VIC4773/2020|EPI_ISL_519159|2020 ···················· ···················· ····················· ···················· ···················· ····················
G 62 hCoV-19/Australia/VIC4774/2020|EPI_ISL_519160|2020 ···················· ···················· ····················· ···················· ···················· ····················
G 63 hCoV-19/Australia/VIC4775/2020|EPI_ISL_519161|2020 ···················· ···················· ····················· ···················· ···················· ····················
G 64 hCoV-19/Australia/VIC4777/2020|EPI_ISL_519163|2020 ···················· ···················· ····················· ···················· ···················· ····················
G 65 hCoV-19/Australia/VIC4778/2020|EPI_ISL_519164|2020 ···················· ···················· ····················· ···················· ···················· ····················
G 66 hCoV-19/Australia/VIC4780/2020|EPI_ISL_519166|2020 ···················· ···················· ····················· ···················· ···················· ····················
G 67 hCoV-19/Australia/VIC4781/2020|EPI_ISL_519167|2020 ···················· ···················· ····················· ···················· ···················· ····················
G 68 hCoV-19/Australia/VIC4782/2020|EPI_ISL_519168|2020 ···················· ···················· ····················· ···················· ···················· ····················
G 69 hCoV-19/Australia/VIC4783/2020|EPI_ISL_519169|2020 ···················· ···················· ····················· ···················· ···················· ····················
G 70 hCoV-19/Australia/VIC4784/2020|EPI_ISL_519170|2020 ···················· ···················· ····················· ···················· ···················· ····················
G 71 hCoV-19/Australia/VIC4785/2020|EPI_ISL_519171|2020 ···················· ···················· ····················· ···················· ···················· ····················
G 72 hCoV-19/Australia/VIC4786/2020|EPI_ISL_519172|2020 ···················· ···················· ····················· ···················· ···················· ····················
G 73 hCoV-19/Australia/VIC4787/2020|EPI_ISL_519173|2020 ···················· ···················· ····················· ···················· ···················· ····················
G 74 hCoV-19/Australia/VIC4788/2020|EPI_ISL_519174|2020 ···················· ···················· ····················· ···················· ···················· ····················
G 75 hCoV-19/Australia/VIC4790/2020|EPI_ISL_519176|2020 ···················· ···················· ····················· ···················· ···················· ····················
G 76 hCoV-19/Australia/VIC4791/2020|EPI_ISL_519177|2020 ···················· ···················· ····················· ···················· ···················· ····················
G 77 hCoV-19/Australia/VIC4792/2020|EPI_ISL_519178|2020 ···················· ···················· ····················· ···················· ···················· ····················
G 78 hCoV-19/Australia/VIC4793/2020|EPI_ISL_519179|2020 ···················· ···················· ····················· ···················· ···················· ····················
G 79 hCoV-19/Australia/VIC5337/2020|EPI_ISL_520100|2020 ···················· ···················· ····················· ···················· ···················· ····················
G 80 hCoV-19/Australia/VIC5338/2020|EPI_ISL_520101|2020 ···················· ···················· ····················· ···················· ···················· ····················
G 81 hCoV-19/Australia/VIC5339/2020|EPI_ISL_520102|2020 ···················· ···················· ····················· ···················· ···················· ····················
G 82 hCoV-19/Australia/VIC5340/2020|EPI_ISL_520103|2020 ···················· ···················· ····················· ···················· ···················· ····················
G 83 hCoV-19/Australia/VIC5341/2020|EPI_ISL_520104|2020 ···················· ···················· ····················· ···················· ···················· ····················
G 84 hCoV-19/Australia/VIC5342/2020|EPI_ISL_520105|2020 ···················· ···················· ····················· ···················· ···················· ····················
G 85 hCoV-19/Australia/VIC5343/2020|EPI_ISL_520106|2020 ···················· ···················· ····················· ···················· ···················· ····················
G 86 hCoV-19/Australia/VIC5344/2020|EPI_ISL_520107|2020 ···················· ···················· ····················· ···················· ···················· ····················
G 87 hCoV-19/Australia/VIC5345/2020|EPI_ISL_520108|2020 ···················· ···················· ····················· ···················· ···················· ····················
G 88 hCoV-19/Australia/VIC5346/2020|EPI_ISL_520109|2020 ···················· ···················· ····················· ···················· ···················· ····················
G 89 hCoV-19/Australia/VIC5347/2020|EPI_ISL_520110|2020 ···················· ···················· ····················· ···················· ···················· ····················
G 90 hCoV-19/Australia/VIC5348/2020|EPI_ISL_520111|2020 ···················· ···················· ····················· ···················· ···················· ····················
G 91 hCoV-19/Australia/VIC5349/2020|EPI_ISL_520112|2020 ···················· ···················· ····················· ···················· ···················· ····················
G 92 hCoV-19/Australia/VIC5350/2020|EPI_ISL_520113|2020 ···················· ···················· ····················· ···················· ···················· ····················
G 93 hCoV-19/Australia/VIC5351/2020|EPI_ISL_520114|2020 ···················· ···················· ····················· ···················· ···················· ····················
G 94 hCoV-19/Australia/VIC5352/2020|EPI_ISL_520115|2020 ···················· ···················· ····················· ···················· ···················· ····················
G 95 hCoV-19/Australia/VIC5353/2020|EPI_ISL_520116|2020 ···················· ···················· ····················· ···················· ···················· ····················
G 96 hCoV-19/Australia/VIC5354/2020|EPI_ISL_520117|2020 ···················· ···················· ····················· ···················· ···················· ····················
G 97 hCoV-19/Australia/VIC5355/2020|EPI_ISL_520118|2020 ···················· ···················· ····················· ···················· ···················· ····················
G 98 hCoV-19/Australia/VIC5356/2020|EPI_ISL_520119|2020 ···················· ···················· ····················· ···················· ···················· ····················
G 99 hCoV-19/Australia/VIC5358/2020|EPI_ISL_520121|2020 ···················· ···················· ····················· ···················· ···················· ····················
G 100 hCoV-19/Australia/VIC5359/2020|EPI_ISL_520122|2020 ···················· ···················· ····················· ···················· ···················· ····················
G 101 hCoV-19/Australia/VIC5360/2020|EPI_ISL_520123|2020 ···················· ···················· ····················· ···················· ···················· ····················
G 102 hCoV-19/Australia/VIC5361/2020|EPI_ISL_520124|2020 ···················· ···················· ····················· ···················· ···················· ····················
G 103 hCoV-19/Australia/VIC5362/2020|EPI_ISL_520125|2020 ···················· ···················· ····················· ···················· ···················· ····················
G 104 hCoV-19/Australia/VIC5363/2020|EPI_ISL_520126|2020 ···················· ···················· ····················· ···················· ···················· ····················
G 105 hCoV-19/Australia/VIC5364/2020|EPI_ISL_520127|2020 ···················· ···················· ····················· ···················· ···················· ····················
G 106 hCoV-19/Australia/VIC5365/2020|EPI_ISL_520128|2020 ···················· ···················· ····················· ···················· ···················· ····················
G 107 hCoV-19/Australia/VIC5366/2020|EPI_ISL_520129|2020 ···················· ···················· ····················· ···················· ···················· ····················
G 108 hCoV-19/Australia/VIC5367/2020|EPI_ISL_520130|2020 ···················· ···················· ····················· ···················· ···················· ····················
G 109 hCoV-19/Australia/VIC5368/2020|EPI_ISL_520131|2020 ···················· ···················· ····················· ···················· ···················· ····················
G 110 hCoV-19/Australia/VIC5369/2020|EPI_ISL_520132|2020 ···················· ···················· ····················· ···················· ···················· ····················

| | | | | | | | | |
|---|---|---|---|---|---|---|---|---|
| G | 111 | hCoV-19/Australia/VIC5370/2020\|EPI_ISL_520133\|2020 | .................... | .................... | ..................... | .................... | .................... | .................... |
| G | 112 | hCoV-19/Australia/VIC5371/2020\|EPI_ISL_520134\|2020 | .................... | .................... | ..................... | .................... | .................... | .................... |
| G | 113 | hCoV-19/Australia/VIC5372/2020\|EPI_ISL_520135\|2020 | .................... | .................... | ..................... | .................... | .................... | .................... |
| G | 114 | hCoV-19/Australia/VIC5373/2020\|EPI_ISL_520136\|2020 | .................... | .................... | ..................... | .................... | .................... | .................... |
| G | 115 | hCoV-19/Australia/VIC5374/2020\|EPI_ISL_520137\|2020 | .................... | .................... | ..................... | .................... | .................... | .................... |
| G | 116 | hCoV-19/Australia/VIC5375/2020\|EPI_ISL_520138\|2020 | .................... | .................... | ..................... | .................... | .................... | .................... |
| G | 117 | hCoV-19/Australia/VIC5376/2020\|EPI_ISL_520139\|2020 | .................... | .................... | ..................... | .................... | .................... | .................... |
| G | 118 | hCoV-19/Australia/VIC5377/2020\|EPI_ISL_520140\|2020 | .................... | .................... | ..................... | .................... | .................... | .................... |
| G | 119 | hCoV-19/Australia/VIC5378/2020\|EPI_ISL_520141\|2020 | .................... | .................... | ..................... | .................... | .................... | .................... |
| G | 120 | hCoV-19/Australia/VIC5379/2020\|EPI_ISL_520142\|2020 | .................... | .................... | ..................... | .................... | .................... | .................... |
| G | 121 | hCoV-19/Australia/VIC5380/2020\|EPI_ISL_520143\|2020 | .................... | .................... | ..................... | .................... | .................... | .................... |
| G | 122 | hCoV-19/Australia/VIC5381/2020\|EPI_ISL_520144\|2020 | .................... | .................... | ..................... | .................... | .................... | .................... |
| G | 123 | hCoV-19/Australia/VIC5382/2020\|EPI_ISL_520145\|2020 | .................... | .................... | ..................... | .................... | .................... | .................... |
| G | 124 | hCoV-19/Australia/VIC5383/2020\|EPI_ISL_520146\|2020 | .................... | .................... | ..................... | .................... | .................... | .................... |
| G | 125 | hCoV-19/Australia/VIC5384/2020\|EPI_ISL_520147\|2020 | .................... | .................... | ..................... | .................... | .................... | .................... |
| G | 126 | hCoV-19/Australia/VIC5385/2020\|EPI_ISL_520148\|2020 | .................... | .................... | ..................... | .................... | .................... | .................... |
| G | 127 | hCoV-19/Australia/VIC5386/2020\|EPI_ISL_520149\|2020 | .................... | .................... | ..................... | .................... | .................... | .................... |
| G | 128 | hCoV-19/Australia/VIC5387/2020\|EPI_ISL_520150\|2020 | .................... | .................... | ..................... | .................... | .................... | .................... |
| G | 129 | hCoV-19/Australia/VIC5388/2020\|EPI_ISL_520151\|2020 | .................... | .................... | ..................... | .................... | .................... | .................... |
| G | 130 | hCoV-19/Australia/VIC5389/2020\|EPI_ISL_520152\|2020 | .................... | .................... | ..................... | .................... | .................... | .................... |
| G | 131 | hCoV-19/Australia/VIC5390/2020\|EPI_ISL_520153\|2020 | .................... | .................... | ..................... | .................... | .................... | .................... |
| G | 132 | hCoV-19/Australia/VIC5391/2020\|EPI_ISL_520154\|2020 | .................... | .................... | ..................... | .................... | .................... | .................... |
| G | 133 | hCoV-19/Australia/VIC5392/2020\|EPI_ISL_520155\|2020 | .................... | .................... | ..................... | .................... | .................... | .................... |
| G | 134 | hCoV-19/Australia/VIC5393/2020\|EPI_ISL_520156\|2020 | .................... | .................... | ..................... | .................... | .................... | .................... |
| G | 135 | hCoV-19/Australia/VIC5394/2020\|EPI_ISL_520157\|2020 | .................... | .................... | ..................... | .................... | .................... | .................... |
| G | 136 | hCoV-19/Australia/VIC5395/2020\|EPI_ISL_520158\|2020 | .................... | .................... | ..................... | .................... | .................... | .................... |
| G | 137 | hCoV-19/Australia/VIC5396/2020\|EPI_ISL_520159\|2020 | .................... | .................... | ..................... | .................... | .................... | .................... |
| G | 138 | hCoV-19/Australia/VIC5397/2020\|EPI_ISL_520160\|2020 | .................... | .................... | ..................... | .................... | .................... | .................... |
| G | 139 | hCoV-19/Australia/VIC5398/2020\|EPI_ISL_520161\|2020 | .................... | .................... | ..................... | .................... | .................... | .................... |
| G | 140 | hCoV-19/Australia/VIC5399/2020\|EPI_ISL_520162\|2020 | .................... | .................... | ..................... | .................... | .................... | .................... |
| G | 141 | hCoV-19/Australia/VIC5400/2020\|EPI_ISL_520163\|2020 | .................... | .................... | ..................... | .................... | .................... | .................... |
| G | 142 | hCoV-19/Australia/VIC5401/2020\|EPI_ISL_520164\|2020 | .................... | .................... | ..................... | .................... | .................... | .................... |
| G | 143 | hCoV-19/Australia/VIC5402/2020\|EPI_ISL_520165\|2020 | .................... | .................... | ..................... | .................... | .................... | .................... |
| G | 144 | hCoV-19/Australia/VIC5403/2020\|EPI_ISL_520166\|2020 | .................... | .................... | ..................... | .................... | .................... | .................... |
| G | 145 | hCoV-19/Australia/VIC5404/2020\|EPI_ISL_520167\|2020 | .................... | .................... | ..................... | .................... | .................... | .................... |
| G | 146 | hCoV-19/Australia/VIC5405/2020\|EPI_ISL_520168\|2020 | .................... | .................... | ..................... | .................... | .................... | .................... |
| G | 147 | hCoV-19/Australia/VIC5406/2020\|EPI_ISL_520169\|2020 | .................... | .................... | ..................... | .................... | .................... | .................... |
| G | 148 | hCoV-19/Australia/VIC6659/2020\|EPI_ISL_521415\|2020 | .................... | .................... | ..................... | .................... | .................... | .................... |
| G | 149 | hCoV-19/Australia/VIC6660/2020\|EPI_ISL_521416\|2020 | .................... | .................... | ..................... | .................... | .................... | .................... |
| G | 150 | hCoV-19/Australia/VIC6662/2020\|EPI_ISL_521418\|2020 | .................... | .................... | ..................... | .................... | .................... | .................... |
| G | 151 | hCoV-19/Australia/VIC6663/2020\|EPI_ISL_521419\|2020 | .................... | .................... | ..................... | .................... | .................... | .................... |
| G | 152 | hCoV-19/Australia/VIC6664/2020\|EPI_ISL_521420\|2020 | .................... | .................... | ..................... | .................... | .................... | .................... |
| G | 153 | hCoV-19/Australia/VIC6665/2020\|EPI_ISL_521421\|2020 | .................... | .................... | ..................... | .................... | .................... | .................... |
| G | 154 | hCoV-19/Australia/VIC6666/2020\|EPI_ISL_521422\|2020 | .................... | .................... | ..................... | .................... | .................... | .................... |
| G | 155 | hCoV-19/Australia/VIC6667/2020\|EPI_ISL_521423\|2020 | .................... | .................... | ..................... | .................... | .................... | .................... |
| G | 156 | hCoV-19/Australia/VIC6668/2020\|EPI_ISL_521424\|2020 | .................... | .................... | ..................... | .................... | .................... | .................... |
| G | 157 | hCoV-19/Australia/VIC6669/2020\|EPI_ISL_521425\|2020 | .................... | .................... | ..................... | .................... | .................... | .................... |
| G | 158 | hCoV-19/Australia/VIC6670/2020\|EPI_ISL_521426\|2020 | .................... | .................... | ..................... | .................... | .................... | .................... |
| G | 159 | hCoV-19/Australia/VIC6671/2020\|EPI_ISL_521427\|2020 | .................... | .................... | ..................... | .................... | .................... | .................... |
| G | 160 | hCoV-19/Australia/VIC6672/2020\|EPI_ISL_521428\|2020 | .................... | .................... | ..................... | .................... | .................... | .................... |
| G | 161 | hCoV-19/Australia/VIC6674/2020\|EPI_ISL_521430\|2020 | .................... | .................... | ..................... | .................... | .................... | .................... |
| G | 162 | hCoV-19/Australia/VIC6675/2020\|EPI_ISL_521431\|2020 | .................... | .................... | ..................... | .................... | .................... | .................... |
| G | 163 | hCoV-19/Australia/VIC6676/2020\|EPI_ISL_521432\|2020 | .................... | .................... | ..................... | .................... | .................... | .................... |
| G | 164 | hCoV-19/Australia/VIC6677/2020\|EPI_ISL_521433\|2020 | .................... | .................... | ..................... | .................... | .................... | .................... |
| G | 165 | hCoV-19/Australia/VIC6678/2020\|EPI_ISL_521434\|2020 | .................... | .................... | ..................... | .................... | .................... | .................... |
| G | 166 | hCoV-19/Australia/VIC6679/2020\|EPI_ISL_521435\|2020 | .................... | .................... | ..................... | .................... | .................... | .................... |
| G | 167 | hCoV-19/Australia/VIC6680/2020\|EPI_ISL_521436\|2020 | .................... | .................... | ..................... | .................... | .................... | .................... |
| G | 168 | hCoV-19/Australia/VIC6682/2020\|EPI_ISL_521438\|2020 | .................... | .................... | ..................... | .................... | .................... | .................... |
| G | 169 | hCoV-19/Australia/VIC6684/2020\|EPI_ISL_521440\|2020 | .................... | .................... | ..................... | .................... | .................... | .................... |
| G | 170 | hCoV-19/Australia/VIC6685/2020\|EPI_ISL_521441\|2020 | .................... | .................... | ..................... | .................... | .................... | .................... |
| G | 171 | hCoV-19/Australia/VIC6686/2020\|EPI_ISL_521442\|2020 | .................... | .................... | ..................... | .................... | .................... | .................... |
| G | 172 | hCoV-19/Australia/VIC6687/2020\|EPI_ISL_521443\|2020 | .................... | .................... | ..................... | .................... | .................... | .................... |
| G | 173 | hCoV-19/Australia/VIC6688/2020\|EPI_ISL_521444\|2020 | .................... | .................... | ..................... | .................... | .................... | .................... |
| G | 174 | hCoV-19/Australia/VIC6689/2020\|EPI_ISL_521445\|2020 | .................... | .................... | ..................... | .................... | .................... | .................... |
| G | 175 | hCoV-19/Australia/VIC6691/2020\|EPI_ISL_521447\|2020 | .................... | .................... | ..................... | .................... | .................... | .................... |
| G | 176 | hCoV-19/Australia/VIC6692/2020\|EPI_ISL_521448\|2020 | .................... | .................... | ..................... | .................... | .................... | .................... |
| G | 177 | hCoV-19/Australia/VIC6693/2020\|EPI_ISL_521449\|2020 | .................... | .................... | ..................... | .................... | .................... | .................... |
| G | 179 | hCoV-19/Australia/VIC2577/2020\|EPI_ISL_521452\|2020 | .................... | .................... | ..................... | .................... | .................... | .................... |
| G | 180 | hCoV-19/Australia/VIC2578/2020\|EPI_ISL_521453\|2020 | .................... | .................... | ..................... | .................... | .................... | .................... |
| G | 181 | hCoV-19/Australia/VIC2579/2020\|EPI_ISL_521454\|2020 | .................... | .................... | ..................... | .................... | .................... | .................... |
| G | 182 | hCoV-19/Australia/VIC2580/2020\|EPI_ISL_521455\|2020 | .................... | .................... | ..................... | .................... | .................... | .................... |
| G | 183 | hCoV-19/Australia/VIC2582/2020\|EPI_ISL_521456\|2020 | .................... | .................... | ..................... | .................... | .................... | .................... |
| G | 184 | hCoV-19/Australia/VIC2583/2020\|EPI_ISL_521457\|2020 | .................... | .................... | ..................... | .................... | .................... | .................... |
| G | 185 | hCoV-19/Australia/VIC2584/2020\|EPI_ISL_521458\|2020 | .................... | .................... | ..................... | .................... | .................... | .................... |
| G | 186 | hCoV-19/Australia/VIC2586/2020\|EPI_ISL_521459\|2020 | .................... | .................... | ..................... | .................... | .................... | .................... |
| G | 187 | hCoV-19/Australia/VIC2587/2020\|EPI_ISL_521460\|2020 | .................... | .................... | ..................... | .................... | .................... | .................... |
| G | 188 | hCoV-19/Australia/VIC2588/2020\|EPI_ISL_521461\|2020 | .................... | .................... | ..................... | .................... | .................... | .................... |
| G | 189 | hCoV-19/Australia/VIC2611/2020\|EPI_ISL_521463\|2020 | .................... | .................... | ..................... | .................... | .................... | .................... |
| G | 190 | hCoV-19/Australia/VIC2612/2020\|EPI_ISL_521464\|2020 | .................... | .................... | ..................... | .................... | .................... | .................... |
| G | 191 | hCoV-19/Australia/VIC2613/2020\|EPI_ISL_521465\|2020 | .................... | .................... | ..................... | .................... | .................... | .................... |

| | | | | | | | | |
|---|---|---|---|---|---|---|---|---|
| G | 192 hCoV-19/Australia/VIC2620/2020\|EPI_ISL_521467\|2020 | ···················· | ···················· | ····················· | ···················· | ···················· | ···················· | |
| G | 193 hCoV-19/Australia/VIC2622/2020\|EPI_ISL_521468\|2020 | ···················· | ···················· | ····················· | ···················· | ···················· | ···················· | |
| G | 194 hCoV-19/Australia/VIC2623/2020\|EPI_ISL_521469\|2020 | ···················· | ···················· | ····················· | ···················· | ···················· | ···················· | |
| G | 195 hCoV-19/Australia/VIC2624/2020\|EPI_ISL_521470\|2020 | ···················· | ···················· | ····················· | ···················· | ···················· | ···················· | |
| G | 196 hCoV-19/Australia/VIC2625/2020\|EPI_ISL_521471\|2020 | ···················· | ···················· | ····················· | ···················· | ···················· | ···················· | |
| G | 197 hCoV-19/Australia/VIC2626/2020\|EPI_ISL_521472\|2020 | ···················· | ···················· | ····················· | ···················· | ···················· | ···················· | |
| G | 198 hCoV-19/Australia/VIC2627/2020\|EPI_ISL_521473\|2020 | ···················· | ···················· | ····················· | ···················· | ···················· | ···················· | |
| G | 199 hCoV-19/Australia/VIC2628/2020\|EPI_ISL_521474\|2020 | ···················· | ···················· | ····················· | ···················· | ···················· | ···················· | |
| G | 200 hCoV-19/Australia/VIC2629/2020\|EPI_ISL_521475\|2020 | ···················· | ···················· | ····················· | ···················· | ···················· | ···················· | |
| G | 201 hCoV-19/Australia/VIC2630/2020\|EPI_ISL_521476\|2020 | ···················· | ···················· | ····················· | ···················· | ···················· | ···················· | |
| G | 202 hCoV-19/Australia/VIC2631/2020\|EPI_ISL_521477\|2020 | ···················· | ···················· | ····················· | ···················· | ···················· | ···················· | |
| G | 203 hCoV-19/Australia/VIC2632/2020\|EPI_ISL_521478\|2020 | ···················· | ···················· | ····················· | ···················· | ···················· | ···················· | |
| G | 204 hCoV-19/Australia/VIC2634/2020\|EPI_ISL_521480\|2020 | ···················· | ···················· | ····················· | ···················· | ···················· | ···················· | |
| G | 205 hCoV-19/Australia/VIC2635/2020\|EPI_ISL_521481\|2020 | ···················· | ···················· | ····················· | ···················· | ···················· | ···················· | |
| G | 206 hCoV-19/Australia/VIC2636/2020\|EPI_ISL_521482\|2020 | ···················· | ···················· | ····················· | ···················· | ···················· | ···················· | |
| G | 207 hCoV-19/Australia/VIC2637/2020\|EPI_ISL_521483\|2020 | ···················· | ···················· | ····················· | ···················· | ···················· | ···················· | |
| G | 209 hCoV-19/Australia/VIC2640/2020\|EPI_ISL_521486\|2020 | ···················· | ···················· | ····················· | ···················· | ···················· | ···················· | |
| G | 210 hCoV-19/Australia/VIC2641/2020\|EPI_ISL_521487\|2020 | ···················· | ···················· | ····················· | ···················· | ···················· | ···················· | |
| G | 211 hCoV-19/Australia/VIC2642/2020\|EPI_ISL_521488\|2020 | ···················· | ···················· | ····················· | ···················· | ···················· | ···················· | |
| G | 212 hCoV-19/Australia/VIC2643/2020\|EPI_ISL_521489\|2020 | ···················· | ···················· | ····················· | ···················· | ···················· | ···················· | |
| G | 213 hCoV-19/Australia/VIC2644/2020\|EPI_ISL_521490\|2020 | ···················· | ···················· | ····················· | ···················· | ···················· | ···················· | |
| G | 214 hCoV-19/Australia/VIC2645/2020\|EPI_ISL_521491\|2020 | ···················· | ···················· | ····················· | ···················· | ···················· | ···················· | |
| G | 215 hCoV-19/Australia/VIC2646/2020\|EPI_ISL_521492\|2020 | ···················· | ···················· | ····················· | ···················· | ···················· | ···················· | |
| G | 216 hCoV-19/Australia/VIC2647/2020\|EPI_ISL_521493\|2020 | ···················· | ···················· | ····················· | ···················· | ···················· | ···················· | |
| G | 217 hCoV-19/Australia/VIC2648/2020\|EPI_ISL_521494\|2020 | ···················· | ···················· | ····················· | ···················· | ···················· | ···················· | |
| G | 218 hCoV-19/Australia/VIC2649/2020\|EPI_ISL_521495\|2020 | ···················· | ···················· | ····················· | ···················· | ···················· | ···················· | |
| G | 219 hCoV-19/Australia/VIC2650/2020\|EPI_ISL_521496\|2020 | ···················· | ···················· | ····················· | ···················· | ···················· | ···················· | |
| G | 220 hCoV-19/Australia/VIC2651/2020\|EPI_ISL_521497\|2020 | ···················· | ···················· | ····················· | ···················· | ···················· | ···················· | |
| G | 221 hCoV-19/Australia/VIC2652/2020\|EPI_ISL_521498\|2020 | ···················· | ···················· | ····················· | ···················· | ···················· | ···················· | |
| G | 223 hCoV-19/Australia/TAS97/2020\|EPI_ISL_522767\|2020-0 | ···················· | ···················· | ····················· | ···················· | NNN················· | ···················· | 1 |
| G | 224 hCoV-19/USA/VA-DCLS-0901/2020\|EPI_ISL_522777\|2020- | ···················· | ···················· | ····················· | ···················· | ···················· | ···················· | |
| G | 225 hCoV-19/USA/VA-DCLS-0902/2020\|EPI_ISL_522778\|2020- | ···················· | ···················· | ····················· | ···················· | ···················· | ···················· | |
| G | 226 hCoV-19/USA/VA-DCLS-0904/2020\|EPI_ISL_522779\|2020- | ···················· | ···················· | ····················· | ···················· | ···················· | ···················· | |
| G | 227 hCoV-19/USA/VA-DCLS-0905/2020\|EPI_ISL_522780\|2020- | ···················· | ···················· | ····················· | ···················· | ···················· | ···················· | |
| G | 228 hCoV-19/USA/VA-DCLS-0908/2020\|EPI_ISL_522781\|2020- | ···················· | ···················· | ····················· | ···················· | ···················· | ···················· | |
| G | 229 hCoV-19/USA/VA-DCLS-0909/2020\|EPI_ISL_522782\|2020- | ···················· | ···················· | ····················· | ···················· | ···················· | ···················· | |
| G | 230 hCoV-19/USA/VA-DCLS-0910/2020\|EPI_ISL_522783\|2020- | ···················· | ···················· | ····················· | ···················· | ···················· | ···················· | |
| G | 231 hCoV-19/USA/VA-DCLS-0912/2020\|EPI_ISL_522784\|2020- | ···················· | ···················· | ····················· | ···················· | ···················· | ···················· | |
| G | 232 hCoV-19/USA/VA-DCLS-0913/2020\|EPI_ISL_522785\|2020- | ···················· | ···················· | ····················· | ···················· | ···················· | ···················· | |
| G | 233 hCoV-19/USA/VA-DCLS-0914/2020\|EPI_ISL_522786\|2020- | ···················· | ···A················ | ····················· | ···················· | ···················· | ···················· | |
| G | 234 hCoV-19/USA/VA-DCLS-0916/2020\|EPI_ISL_522787\|2020- | ···················· | ···················· | ····················· | ···················· | ···················· | ···················· | |
| G | 235 hCoV-19/USA/VA-DCLS-0917/2020\|EPI_ISL_522788\|2020- | ···················· | ···················· | ····················· | ···················· | ···················· | ···················· | |
| G | 236 hCoV-19/USA/VA-DCLS-0918/2020\|EPI_ISL_522789\|2020- | ···················· | ···················· | ····················· | ···················· | ···················· | ···················· | |
| G | 237 hCoV-19/USA/VA-DCLS-0919/2020\|EPI_ISL_522790\|2020- | ···················· | ···················· | ····················· | ···················· | ···················· | ···················· | |
| G | 238 hCoV-19/USA/VA-DCLS-0920/2020\|EPI_ISL_522791\|2020- | ···················· | ···················· | ····················· | ···················· | ···················· | ···················· | |
| G | 239 hCoV-19/USA/VA-DCLS-0922/2020\|EPI_ISL_522792\|2020- | ···················· | ···················· | ····················· | ···················· | ···················· | ···················· | |
| G | 240 hCoV-19/USA/VA-DCLS-0923/2020\|EPI_ISL_522793\|2020- | ···················· | ···················· | ····················· | ···················· | ···················· | ···················· | |
| G | 241 hCoV-19/USA/VA-DCLS-0924/2020\|EPI_ISL_522794\|2020- | ···················· | ···················· | ····················· | ···················· | ···················· | ···················· | |
| G | 242 hCoV-19/USA/VA-DCLS-0926/2020\|EPI_ISL_522795\|2020- | ···················· | ···················· | ····················· | ···················· | ···················· | ···················· | |
| G | 243 hCoV-19/USA/VA-DCLS-0928/2020\|EPI_ISL_522796\|2020- | ···················· | ···················· | ····················· | ···················· | ···················· | ···················· | |
| G | 244 hCoV-19/USA/VA-DCLS-0930/2020\|EPI_ISL_522797\|2020- | ···················· | ···················· | ····················· | ···················· | ···················· | ···················· | |
| G | 245 hCoV-19/USA/VA-DCLS-0933/2020\|EPI_ISL_522798\|2020- | ···················· | ···················· | ····················· | ···················· | ···················· | ···················· | |
| G | 246 hCoV-19/USA/VA-DCLS-0934/2020\|EPI_ISL_522799\|2020- | ···················· | ···················· | ····················· | ···················· | ···················· | ···················· | |
| G | 247 hCoV-19/England/ALDP-97C385/2020\|EPI_ISL_526301\|20 | ···················· | ···················· | ····················· | ···················· | ···················· | ···················· | |
| G | 248 hCoV-19/England/ALDP-97C929/2020\|EPI_ISL_526304\|20 | ···················· | ···················· | ····················· | ···················· | ···················· | ···················· | |
| G | 249 hCoV-19/England/ALDP-97C947/2020\|EPI_ISL_526305\|20 | ···················· | ···················· | ····················· | ···················· | ···················· | ···················· | |
| G | 250 hCoV-19/USA/MN-MDH-1565/2020\|EPI_ISL_527630\|2020-0 | ···················· | ···················· | ····················· | ···················· | ···················· | ···················· | |
| G | 251 hCoV-19/USA/MN-MDH-1566/2020\|EPI_ISL_527631\|2020-0 | ···················· | ···················· | ····················· | ···················· | ···················· | ···················· | |
| G | 252 hCoV-19/USA/AR-CDC-4205/2020\|EPI_ISL_527632\|2020-0 | ···················· | ···················· | ····················· | ···················· | ···················· | ···················· | |
| G | 253 hCoV-19/USA/AR-CDC-4213/2020\|EPI_ISL_527635\|2020-0 | ···················· | ···················· | ····················· | ···················· | ···················· | ···················· | |
| G | 254 hCoV-19/USA/AR-CDC-4218/2020\|EPI_ISL_527636\|2020-0 | ···················· | ···················· | ····················· | ···················· | ···················· | ···················· | |
| G | 255 hCoV-19/USA/AR-CDC-4219/2020\|EPI_ISL_527637\|2020-0 | ···················· | ···················· | ····················· | ···················· | ···················· | ···················· | |
| G | 256 hCoV-19/USA/AR-CDC-4220/2020\|EPI_ISL_527638\|2020-0 | ···················· | ···················· | ····················· | ···················· | ···················· | ···················· | |
| G | 257 hCoV-19/Israel/CVL1133/2020\|EPI_ISL_516903\|2020-06 | ···················· | ···················· | ····················· | ···················· | ···················· | ···················· | |
| G | 258 hCoV-19/Israel/CVL1196/2020\|EPI_ISL_516905\|2020-06 | ···················· | ···················· | ····················· | ···················· | ···················· | ···················· | |
| G | 259 hCoV-19/Israel/CVL1198/2020\|EPI_ISL_516906\|2020-06 | ···················· | ···A················ | ····················· | ···················· | ···················· | ···················· | |
| G | 260 hCoV-19/Israel/CVL4572/2020\|EPI_ISL_516913\|2020-06 | ···················· | ·A·················· | ····················· | ···················· | ···················· | ···················· | |
| G | 261 hCoV-19/Israel/CVL4577/2020\|EPI_ISL_516915\|2020-06 | ···················· | ·A·················· | ····················· | ···················· | ···················· | ···················· | |
| G | 262 hCoV-19/Israel/CVL5421/2020\|EPI_ISL_516917\|2020-06 | ···················· | ···················· | ····················· | ···················· | ···················· | ···················· | |
| G | 263 hCoV-19/Israel/CVL5786/2020\|EPI_ISL_516919\|2020-06 | ···················· | ·A·················· | ····················· | ···················· | ···················· | ···················· | |
| G | 264 hCoV-19/Israel/CVL5972/2020\|EPI_ISL_516920\|2020-06 | ···················· | ···················· | ····················· | ···················· | ···················· | ···················· | |
| G | 265 hCoV-19/Israel/CVL6027/2020\|EPI_ISL_516921\|2020-06 | ···················· | ···················· | ····················· | ···················· | ···················· | ··············G····· | |
| G | 266 hCoV-19/Montenegro/ChVir-1265622002/2020\|EPI_ISL_5 | ···················· | ···················· | ····················· | ···················· | ···················· | ···················· | |
| G | 267 hCoV-19/Montenegro/ChVir-1267392002/2020\|EPI_ISL_5 | ···················· | ···················· | ····················· | ···················· | ···················· | ···················· | |
| G | 268 hCoV-19/Montenegro/ChVir-1268662002/2020\|EPI_ISL_5 | ···················· | ···················· | ····················· | ···················· | ···················· | ···················· | |
| G | 269 hCoV-19/Montenegro/ChVir-1793232002/2020\|EPI_ISL_5 | ···················· | ···················· | ····················· | ···················· | ···················· | ···················· | |
| G | 270 hCoV-19/Montenegro/ChVir-1794332002/2020\|EPI_ISL_5 | ···················· | ···················· | ····················· | ···················· | ···················· | ···················· | |
| G | 271 hCoV-19/Montenegro/ChVir-1796572002/2020\|EPI_ISL_5 | ···················· | ···················· | ····················· | ···················· | ···················· | ···················· | |
| G | 272 hCoV-19/Montenegro/ChVir-1800902002/2020\|EPI_ISL_5 | ···················· | ···················· | ····················· | ···················· | ···················· | ···················· | |
| G | 273 hCoV-19/Montenegro/ChVir-1800932002/2020\|EPI_ISL_5 | ···················· | ···················· | ····················· | ···················· | ···················· | ···················· | |

| | | | | | | | |
|---|---|---|---|---|---|---|---|
| G | 274 hCoV-19/Montenegro/ChVir-1800942002/2020\|EPI_ISL_5 | ···················· | ···················· | ····················· | ···················· | ···················· | ···················· |
| G | 275 hCoV-19/Montenegro/ChVir-1804702002/2020\|EPI_ISL_5 | ···················· | ···················· | ····················· | ···················· | ···················· | ···················· |
| G | 276 hCoV-19/Montenegro/ChVir-1807082002/2020\|EPI_ISL_5 | ···················· | ···················· | ····················· | ···················· | ···················· | ···················· |
| G | 277 hCoV-19/Montenegro/ChVir-1807132002/2020\|EPI_ISL_5 | ···················· | ···················· | ····················· | ···················· | ···················· | ···················· |
| G | 278 hCoV-19/Moldova/ICGEB_MD1/2020\|EPI_ISL_516934\|2020 | ···················· | ···················· | ····················· | ···················· | ···················· | ···················· |
| G | 279 hCoV-19/Moldova/ICGEB_MD6/2020\|EPI_ISL_516938\|2020 | ···················· | ···················· | ····················· | ···················· | ···················· | ···················· |
| G | 280 hCoV-19/India/UP-NBRI-N11-TCR316/2020\|EPI_ISL_5169 | ···················· | ···················· | ····················· | ···················· | ···················· | ···················· |
| G | 281 hCoV-19/India/UP-NBRI-N13-TCR323/2020\|EPI_ISL_5169 | ···················· | ···················· | ····················· | ···················· | ···················· | ···················· |
| G | 282 hCoV-19/India/UP-NBRI-N14-TCR324/2020\|EPI_ISL_5169 | ···················· | ···················· | ····················· | ···················· | ···················· | ···················· |
| G | 283 hCoV-19/India/UP-NBRI-N17-TCP057/2020\|EPI_ISL_5169 | ···················· | ···················· | ····················· | ···················· | ···················· | ···················· |
| G | 284 hCoV-19/India/UP-NBRI-N19-TCP645/2020\|EPI_ISL_5169 | ···················· | ···················· | ····················· | ···················· | ···················· | ···················· |
| G | 285 hCoV-19/India/UP-NBRI-N20-TDU302/2020\|EPI_ISL_5169 | ···················· | ···················· | ····················· | ···················· | ···················· | ···················· |
| G | 286 hCoV-19/India/UP-NBRI-N40-TCQ729/2020\|EPI_ISL_5169 | ···················· | ···················· | ····················· | ···················· | ···················· | ···················· |
| G | 287 hCoV-19/India/UP-NBRI-N46-TCQ289/2020\|EPI_ISL_5169 | ···················· | ···················· | ····················· | ···················· | ···················· | ···················· |
| G | 288 hCoV-19/India/UP-NBRI-N48-TEI094/2020\|EPI_ISL_5169 | ···················· | ·A·················· | ····················· | ···················· | ···················· | ··A················· |
| G | 289 hCoV-19/India/UP-NBRI-N49-TEI217/2020\|EPI_ISL_5169 | ···················· | ·A·················· | ····················· | ···················· | ···················· | ··A················· |
| G | 290 hCoV-19/India/UP-NBRI-N52-TEH623/2020\|EPI_ISL_5169 | ···················· | ·A·················· | ····················· | ···················· | ···················· | ··A················· |
| G | 291 hCoV-19/India/UP-NBRI-N53-TEH628/2020\|EPI_ISL_5169 | ···················· | ·A·················· | ····················· | ···················· | ···················· | ··A················· |
| G | 292 hCoV-19/India/UP-NBRI-N54-TEH857/2020\|EPI_ISL_5169 | ···················· | ·A·················· | ····················· | ···················· | ···················· | ··A················· |
| G | 293 hCoV-19/India/UP-NBRI-N55-TEH861/2020\|EPI_ISL_5169 | ···················· | ·A·················· | ····················· | ···················· | ···················· | ··A················· |
| G | 294 hCoV-19/India/UP-NBRI-N56-TEH862/2020\|EPI_ISL_5169 | ···················· | ·A·················· | ····················· | ···················· | ···················· | ··A················· |
| G | 295 hCoV-19/India/UP-NBRI-N58-TEG811/2020\|EPI_ISL_5169 | ···················· | ·A·················· | ····················· | ···················· | ···················· | ··A················· |
| G | 296 hCoV-19/Peru/ANC-INS-066/2020\|EPI_ISL_516987\|2020- | ···················· | ···················· | ····················· | ···················· | ···················· | ···················· |
| G | 297 hCoV-19/Slovakia/ChVir-1996/2020\|EPI_ISL_516988\|20 | ···················· | ···················· | ····················· | ···················· | ···················· | ···················· |
| G | 298 hCoV-19/Slovakia/ChVir-1998/2020\|EPI_ISL_516989\|20 | ···················· | ···················· | ····················· | ···················· | ···················· | ···················· |
| G | 299 hCoV-19/Australia/VIC4794/2020\|EPI_ISL_519180\|2020 | ···················· | ···················· | ····················· | ···················· | ···················· | ···················· |
| G | 300 hCoV-19/Australia/VIC4795/2020\|EPI_ISL_519181\|2020 | ···················· | ···················· | ····················· | ···················· | ···················· | ···················· |
| G | 301 hCoV-19/Australia/VIC4796/2020\|EPI_ISL_519182\|2020 | ···················· | ···················· | ····················· | ···················· | ···················· | ···················· |
| G | 302 hCoV-19/Australia/VIC4797/2020\|EPI_ISL_519183\|2020 | ···················· | ···················· | ····················· | ···················· | ···················· | ···················· |
| G | 303 hCoV-19/Australia/VIC4798/2020\|EPI_ISL_519184\|2020 | ···················· | ···················· | ····················· | ···················· | ···················· | ···················· |
| G | 304 hCoV-19/Australia/VIC4799/2020\|EPI_ISL_519185\|2020 | ···················· | ···················· | ····················· | ···················· | ···················· | ···················· |
| G | 305 hCoV-19/Australia/VIC4802/2020\|EPI_ISL_519187\|2020 | ···················· | ···················· | ····················· | ···················· | ···················· | ···················· |
| G | 306 hCoV-19/Australia/VIC4804/2020\|EPI_ISL_519189\|2020 | ···················· | ···················· | ····················· | ···················· | ···················· | ···················· |
| G | 307 hCoV-19/Australia/VIC4805/2020\|EPI_ISL_519190\|2020 | ···················· | ···················· | ····················· | ···················· | ···················· | ···················· |
| G | 308 hCoV-19/Australia/VIC4806/2020\|EPI_ISL_519191\|2020 | ···················· | ···················· | ····················· | ···················· | ···················· | ···················· |
| G | 309 hCoV-19/Australia/VIC4808/2020\|EPI_ISL_519193\|2020 | ···················· | ···················· | ····················· | ···················· | ···················· | ···················· |
| G | 310 hCoV-19/Australia/VIC4809/2020\|EPI_ISL_519194\|2020 | ···················· | ···················· | ····················· | ···················· | ···················· | ···················· |
| G | 311 hCoV-19/Australia/VIC4810/2020\|EPI_ISL_519195\|2020 | ···················· | ···················· | ····················· | ···················· | ···················· | ···················· |
| G | 312 hCoV-19/Australia/VIC4811/2020\|EPI_ISL_519196\|2020 | ···················· | ···················· | ····················· | ···················· | ···················· | ···················· |
| G | 313 hCoV-19/Australia/VIC4813/2020\|EPI_ISL_519198\|2020 | ···················· | ···················· | ····················· | ···················· | ···················· | ···················· |
| G | 314 hCoV-19/Australia/VIC4814/2020\|EPI_ISL_519199\|2020 | ···················· | ···················· | ····················· | ···················· | ···················· | ···················· |
| G | 315 hCoV-19/Australia/VIC5407/2020\|EPI_ISL_520170\|2020 | ···················· | ···················· | ····················· | ···················· | ···················· | ···················· |
| G | 316 hCoV-19/Australia/VIC5408/2020\|EPI_ISL_520171\|2020 | ···················· | ···················· | ····················· | ···················· | ···················· | ···················· |
| G | 317 hCoV-19/Australia/VIC5409/2020\|EPI_ISL_520172\|2020 | ···················· | ···················· | ····················· | ···················· | ···················· | ···················· |
| G | 318 hCoV-19/Australia/VIC5410/2020\|EPI_ISL_520173\|2020 | ···················· | ···················· | ····················· | ···················· | ···················· | ···················· |
| G | 319 hCoV-19/Australia/VIC5411/2020\|EPI_ISL_520174\|2020 | ···················· | ···················· | ····················· | ···················· | ···················· | ···················· |
| G | 320 hCoV-19/Australia/VIC5412/2020\|EPI_ISL_520175\|2020 | ···················· | ···················· | ····················· | ···················· | ···················· | ···················· |
| G | 321 hCoV-19/Australia/VIC5413/2020\|EPI_ISL_520176\|2020 | ···················· | ···················· | ····················· | ···················· | ···················· | ···················· |
| G | 322 hCoV-19/Australia/VIC5414/2020\|EPI_ISL_520177\|2020 | ···················· | ···················· | ····················· | ···················· | ···················· | ···················· |
| G | 323 hCoV-19/Australia/VIC5415/2020\|EPI_ISL_520178\|2020 | ···················· | ···················· | ····················· | ···················· | ···················· | ···················· |
| G | 324 hCoV-19/Australia/VIC5417/2020\|EPI_ISL_520179\|2020 | ···················· | ···················· | ····················· | ···················· | ···················· | ···················· |
| G | 325 hCoV-19/Australia/VIC5418/2020\|EPI_ISL_520180\|2020 | ···················· | ···················· | ····················· | ···················· | ···················· | ···················· |
| G | 326 hCoV-19/Australia/VIC5432/2020\|EPI_ISL_520183\|2020 | ···················· | ···················· | ····················· | ···················· | ···················· | ···················· |
| G | 327 hCoV-19/Australia/VIC5433/2020\|EPI_ISL_520184\|2020 | ···················· | ···················· | ····················· | ···················· | ···················· | ···················· |
| G | 328 hCoV-19/Australia/VIC5434/2020\|EPI_ISL_520185\|2020 | ···················· | ···················· | ····················· | ···················· | ···················· | ···················· |
| G | 329 hCoV-19/Australia/VIC5435/2020\|EPI_ISL_520186\|2020 | ···················· | ···················· | ····················· | ···················· | ···················· | ···················· |
| G | 330 hCoV-19/Australia/VIC5437/2020\|EPI_ISL_520188\|2020 | ···················· | ···················· | ····················· | ···················· | ···················· | ···················· |
| G | 331 hCoV-19/Australia/VIC5438/2020\|EPI_ISL_520189\|2020 | ···················· | ···················· | ····················· | ···················· | ···················· | ···················· |
| G | 332 hCoV-19/Australia/VIC5439/2020\|EPI_ISL_520190\|2020 | ···················· | ···················· | ····················· | ···················· | ···················· | ···················· |
| G | 333 hCoV-19/Australia/VIC5441/2020\|EPI_ISL_520192\|2020 | ···················· | ···················· | ····················· | ···················· | ···················· | ···················· |
| G | 334 hCoV-19/Australia/VIC5442/2020\|EPI_ISL_520193\|2020 | ···················· | ···················· | ····················· | ···················· | ···················· | ···················· |
| G | 335 hCoV-19/Australia/VIC5443/2020\|EPI_ISL_520194\|2020 | ···················· | ···················· | ····················· | ···················· | ···················· | ···················· |
| G | 336 hCoV-19/Australia/VIC5444/2020\|EPI_ISL_520195\|2020 | ···················· | ···················· | ····················· | ···················· | ···················· | ···················· |
| G | 337 hCoV-19/Australia/VIC5445/2020\|EPI_ISL_520196\|2020 | ···················· | ···················· | ····················· | ···················· | ···················· | ···················· |
| G | 338 hCoV-19/Australia/VIC5446/2020\|EPI_ISL_520197\|2020 | ···················· | ···················· | ····················· | ···················· | ···················· | ···················· |
| G | 339 hCoV-19/Australia/VIC5447/2020\|EPI_ISL_520198\|2020 | ···················· | ···················· | ····················· | ···················· | ···················· | ···················· |
| G | 340 hCoV-19/Australia/VIC5448/2020\|EPI_ISL_520199\|2020 | ···················· | ···················· | ····················· | ···················· | ···················· | ···················· |
| G | 341 hCoV-19/USA/UT-UPHL-2008896/2020\|EPI_ISL_525010\|20 | ···················· | ···················· | ····················· | ···················· | ···················· | ···················· |
| G | 342 hCoV-19/USA/UT-UPHL-2008897/2020\|EPI_ISL_525011\|20 | ···················· | ···················· | ····················· | ···················· | ···················· | ···················· |
| G | 343 hCoV-19/USA/UT-UPHL-2008901/2020\|EPI_ISL_525014\|20 | ···················· | ·A·················· | ····················· | ···················· | ···················· | ···················· |
| G | 344 hCoV-19/USA/UT-UPHL-2008906/2020\|EPI_ISL_525016\|20 | ···················· | ···················· | ····················· | ···················· | ···················· | ···················· |
| G | 345 hCoV-19/USA/UT-UPHL-2008914/2020\|EPI_ISL_525020\|20 | ···················· | ···················· | ····················· | ···················· | ···················· | ···················· |
| G | 346 hCoV-19/USA/UT-UPHL-2008915/2020\|EPI_ISL_525021\|20 | ···················· | ···················· | ····················· | ···················· | ···················· | ···················· |
| G | 347 hCoV-19/USA/UT-UPHL-2008916/2020\|EPI_ISL_525022\|20 | ···················· | ·A·················· | ····················· | ···················· | ···················· | ···················· |
| G | 348 hCoV-19/USA/UT-UPHL-2008917/2020\|EPI_ISL_525023\|20 | ···················· | ·A·················· | ····················· | ···················· | ···················· | ···················· |
| G | 349 hCoV-19/USA/UT-UPHL-2008919/2020\|EPI_ISL_525025\|20 | ···················· | ···················· | ····················· | ···················· | ···················· | ···················· |
| G | 350 hCoV-19/USA/UT-UPHL-2008921/2020\|EPI_ISL_525027\|20 | ···················· | ···················· | ····················· | ···················· | ···················· | ···················· |
| G | 351 hCoV-19/USA/UT-UPHL-2008922/2020\|EPI_ISL_525028\|20 | ···················· | ···················· | ····················· | ···················· | ···················· | ···················· |
| G | 352 hCoV-19/USA/UT-UPHL-2008923/2020\|EPI_ISL_525029\|20 | ···················· | ···················· | ····················· | ···················· | ···················· | ···················· |
| G | 353 hCoV-19/USA/UT-UPHL-2008924/2020\|EPI_ISL_525030\|20 | ···················· | ·A·················· | ····················· | ···················· | ···················· | ···················· |

| | | | | | | | | |
|---|---|---|---|---|---|---|---|---|
| G | 354 hCoV-19/USA/UT-UPHL-2008927/2020\|EPI_ISL_525032\|20 | ···················· | ···················· | ···················· | ···················· | ···················· | ···················· | |
| G | 355 hCoV-19/USA/UT-UPHL-2008928/2020\|EPI_ISL_525033\|20 | ···················· | ·A·················· | ···················· | ···················· | ···················· | ···················· | |
| G | 356 hCoV-19/USA/UT-UPHL-2008929/2020\|EPI_ISL_525034\|20 | ···················· | ···················· | ···················· | ···················· | ···················· | ···················· | |
| G | 357 hCoV-19/USA/UT-UPHL-2008931/2020\|EPI_ISL_525036\|20 | ···················· | ···················· | ···················· | ···················· | ···················· | ···················· | |
| G | 358 hCoV-19/USA/UT-UPHL-2008932/2020\|EPI_ISL_525037\|20 | ···················· | ···················· | ···················· | ···················· | ···················· | ···················· | |
| G | 359 hCoV-19/USA/UT-UPHL-2008936/2020\|EPI_ISL_525041\|20 | ···················· | ···················· | ···················· | ···················· | ···················· | ···················· | |
| G | 360 hCoV-19/USA/UT-UPHL-2008937/2020\|EPI_ISL_525042\|20 | ···················· | ···················· | ···················· | ···················· | ···················· | ···················· | |
| G | 361 hCoV-19/USA/UT-UPHL-2008938/2020\|EPI_ISL_525043\|20 | ···················· | ···················· | ···················· | ···················· | ···················· | ···················· | |
| G | 362 hCoV-19/USA/UT-UPHL-2008939/2020\|EPI_ISL_525044\|20 | ···················· | ···················· | ···················· | ···················· | ···················· | ···················· | |
| G | 363 hCoV-19/USA/UT-UPHL-2008941/2020\|EPI_ISL_525045\|20 | ···················· | ···················· | ···················· | ···················· | ·········A·········· | ···················· | |
| G | 364 hCoV-19/USA/UT-UPHL-2009032/2020\|EPI_ISL_525048\|20 | ···················· | ···················· | ···················· | ···················· | ···C················ | ···················· | |
| G | 365 hCoV-19/USA/UT-UPHL-2009033/2020\|EPI_ISL_525049\|20 | ···················· | ···················· | ···················· | ···················· | ···················· | ···················· | |
| G | 366 hCoV-19/USA/UT-UPHL-2009035/2020\|EPI_ISL_525050\|20 | ···················· | ···················· | ···················· | ···················· | ···················· | ···················· | |
| G | 367 hCoV-19/USA/UT-UPHL-2009037/2020\|EPI_ISL_525052\|20 | ···················· | ···················· | ···················· | ···················· | ···················· | ···················· | |
| G | 368 hCoV-19/USA/UT-UPHL-2009044/2020\|EPI_ISL_525058\|20 | ···················· | ···················· | ···················· | ···················· | ···················· | ···················· | |
| G | 369 hCoV-19/England/ALDP-97C367/2020\|EPI_ISL_526300\|20 | ···················· | ···················· | ···················· | ···················· | ···················· | ···················· | |
| G | 370 hCoV-19/England/ALDP-97DCC5/2020\|EPI_ISL_526330\|20 | ···················· | ···················· | ···················· | ···················· | ···················· | ···················· | |
| G | 371 hCoV-19/England/ALDP-97DCD4/2020\|EPI_ISL_526331\|20 | ···················· | ···················· | ···················· | ···················· | ···················· | ···················· | |
| G | 372 hCoV-19/England/ALDP-97DD4A/2020\|EPI_ISL_526332\|20 | ···················· | ···················· | ···················· | ···················· | ···················· | ···················· | |
| G | 373 hCoV-19/England/ALDP-97FB5D/2020\|EPI_ISL_526340\|20 | ···················· | ···················· | ···················· | ···················· | ···················· | ···················· | |
| G | 374 hCoV-19/England/ALDP-97FE27/2020\|EPI_ISL_526341\|20 | ···················· | ···················· | ···················· | ···················· | ···················· | ···················· | |
| G | 375 hCoV-19/England/ALDP-97FE45/2020\|EPI_ISL_526343\|20 | ···················· | ···················· | ···················· | ···················· | ···················· | ···················· | |
| G | 376 hCoV-19/England/ALDP-9802AF/2020\|EPI_ISL_526346\|20 | ···················· | ···················· | ···················· | ···················· | ···················· | ···················· | |
| G | 377 hCoV-19/England/ALDP-980579/2020\|EPI_ISL_526348\|20 | ···················· | ···················· | ···················· | ···················· | ···················· | ···················· | |
| G | 378 hCoV-19/England/ALDP-9805A6/2020\|EPI_ISL_526350\|20 | ···················· | ·A·················· | ···················· | ···················· | ···················· | ··A················· | |
| G | 379 hCoV-19/England/ALDP-980BB3/2020\|EPI_ISL_526351\|20 | ···················· | ···················· | ···················· | ···················· | ···················· | ···················· | |
| G | 380 hCoV-19/England/ALDP-981059/2020\|EPI_ISL_526352\|20 | ···················· | ···················· | ···················· | ···················· | ···················· | ···················· | |
| G | 381 hCoV-19/England/ALDP-98150F/2020\|EPI_ISL_526353\|20 | ···················· | ···················· | ···················· | ···················· | ···················· | ···················· | |
| G | 382 hCoV-19/England/ALDP-9819E5/2020\|EPI_ISL_526355\|20 | ···················· | ·A·················· | ···················· | ···················· | ···················· | ··A················· | |
| G | 383 hCoV-19/England/ALDP-9819F4/2020\|EPI_ISL_526356\|20 | ···················· | ···················· | ···················· | ···················· | ···················· | ···················· | |
| G | 384 hCoV-19/England/ALDP-981A1F/2020\|EPI_ISL_526358\|20 | ···················· | ···················· | ···················· | ···················· | ···················· | ···················· | |
| G | 385 hCoV-19/England/ALDP-982207/2020\|EPI_ISL_526360\|20 | ···················· | ·A·················· | ···················· | ···················· | ···················· | ··A················· | |
| G | 386 hCoV-19/England/ALDP-9823F5/2020\|EPI_ISL_526361\|20 | ···················· | ···················· | ···················· | ···················· | ···················· | ···················· | |
| G | 387 hCoV-19/England/ALDP-98243E/2020\|EPI_ISL_526362\|20 | ···················· | ···················· | ···················· | ···················· | ···················· | ···················· | |
| G | 388 hCoV-19/England/ALDP-9826CF/2020\|EPI_ISL_526366\|20 | ···················· | ···················· | ···················· | ···················· | ···················· | ···················· | |
| G | 389 hCoV-19/England/LIVE-A935B/2020\|EPI_ISL_526368\|202 | ···················· | ···················· | ···················· | ···················· | ···················· | ···················· | |
| G | 390 hCoV-19/England/LIVE-A9397/2020\|EPI_ISL_526369\|202 | ···················· | ···················· | ···················· | ···················· | ···················· | ···················· | |
| G | 391 hCoV-19/England/LIVE-A93A6/2020\|EPI_ISL_526370\|202 | ···················· | ···················· | ···················· | ···················· | ···················· | ···················· | |
| G | 392 hCoV-19/England/LIVE-A93C4/2020\|EPI_ISL_526371\|202 | ···················· | ···················· | ···················· | ···················· | ···················· | ···················· | |
| G | 394 hCoV-19/England/LIVE-A941C/2020\|EPI_ISL_526373\|202 | ···················· | ···················· | ···················· | ···················· | ···················· | ···················· | |
| G | 395 hCoV-19/England/LIVE-A943A/2020\|EPI_ISL_526374\|202 | ···················· | ···················· | ···················· | ···················· | ···················· | ···················· | |
| G | 396 hCoV-19/England/LIVE-A9485/2020\|EPI_ISL_526375\|202 | ···················· | ···················· | ···················· | ···················· | ···················· | ···················· | |
| G | 397 hCoV-19/England/LIVE-A94D0/2020\|EPI_ISL_526376\|202 | ···················· | ···················· | ···················· | ···················· | ···················· | ···················· | |
| G | 398 hCoV-19/England/LIVE-A94EF/2020\|EPI_ISL_526377\|202 | ···················· | ···················· | ···················· | ···················· | ···················· | ···················· | |
| G | 399 hCoV-19/England/LIVE-A9528/2020\|EPI_ISL_526378\|202 | ···················· | ···················· | ···················· | ···················· | ···················· | ···················· | |
| G | 400 hCoV-19/England/LIVE-A9564/2020\|EPI_ISL_526379\|202 | ···················· | ···················· | ···················· | ···················· | ···················· | ···················· | |
| G | 401 hCoV-19/England/LIVE-A95EC/2020\|EPI_ISL_526380\|202 | ···················· | ···M················ | ···················· | ···················· | ···················· | ···················· | |
| G | 402 hCoV-19/England/LIVE-A9616/2020\|EPI_ISL_526381\|202 | ···················· | ···················· | ···················· | ···················· | ···················· | ···················· | |
| G | 403 hCoV-19/England/LIVE-A9634/2020\|EPI_ISL_526382\|202 | ···················· | ···················· | ···················· | ···················· | ···················· | ···················· | |
| G | 405 hCoV-19/England/LIVE-A9670/2020\|EPI_ISL_526384\|202 | ···················· | ···················· | ···················· | ···················· | ···················· | ···················· | |
| G | 406 hCoV-19/England/LIVE-A968F/2020\|EPI_ISL_526385\|202 | ···················· | ···M················ | ···················· | ···················· | ···················· | ···················· | |
| G | 407 hCoV-19/England/LIVE-A9713/2020\|EPI_ISL_526386\|202 | ···················· | ···················· | ···················· | ···················· | ···················· | ···················· | |
| G | 408 hCoV-19/England/LIVE-A9722/2020\|EPI_ISL_526387\|202 | ···················· | ···················· | ···················· | ···················· | ···················· | ···················· | |
| G | 409 hCoV-19/England/LIVE-A9731/2020\|EPI_ISL_526388\|202 | ···················· | ···················· | ···················· | ···················· | ···················· | ···················· | |
| G | 410 hCoV-19/England/LIVE-A9740/2020\|EPI_ISL_526389\|202 | ···················· | ···················· | ···················· | ···················· | ···················· | ···················· | |
| G | 411 hCoV-19/USA/MN-MDH-1535/2020\|EPI_ISL_527600\|2020-0 | ···················· | ·A·················· | ···················· | ···················· | ···················· | ···················· | |
| G | 412 hCoV-19/USA/MN-MDH-1536/2020\|EPI_ISL_527601\|2020-0 | ···················· | ·A·················· | ···················· | ···················· | ···················· | ···················· | |
| G | 413 hCoV-19/USA/MN-MDH-1537/2020\|EPI_ISL_527602\|2020-0 | ···················· | ·A·················· | ···················· | ···················· | ···················· | ···················· | |
| G | 414 hCoV-19/USA/MN-MDH-1538/2020\|EPI_ISL_527603\|2020-0 | ···················· | ·A·················· | ···················· | ···················· | ···················· | ···················· | |
| G | 415 hCoV-19/USA/MN-MDH-1539/2020\|EPI_ISL_527604\|2020-0 | ···················· | ·A·················· | ···················· | ···················· | ···················· | ···················· | |
| G | 416 hCoV-19/USA/MN-MDH-1540/2020\|EPI_ISL_527605\|2020-0 | ···················· | ·A·················· | ···················· | ···················· | ···················· | ···················· | |
| G | 417 hCoV-19/USA/MN-MDH-1541/2020\|EPI_ISL_527606\|2020-0 | ···················· | ·A·················· | ···················· | ···················· | ···················· | ···················· | |
| G | 418 hCoV-19/USA/MN-MDH-1542/2020\|EPI_ISL_527607\|2020-0 | ···················· | ·A·················· | ···················· | ···················· | ···················· | ···················· | |
| G | 419 hCoV-19/USA/MN-MDH-1543/2020\|EPI_ISL_527608\|2020-0 | ···················· | ·A·················· | ···················· | ···················· | ·NNN················ | ···················· | 1 |
| G | 420 hCoV-19/USA/MN-MDH-1544/2020\|EPI_ISL_527609\|2020-0 | ···················· | ·A·················· | ···················· | ···················· | ···················· | ···················· | |
| G | 421 hCoV-19/USA/MN-MDH-1545/2020\|EPI_ISL_527610\|2020-0 | ···················· | ·A·················· | ···················· | ···················· | ···················· | ···················· | |
| G | 422 hCoV-19/USA/MN-MDH-1546/2020\|EPI_ISL_527611\|2020-0 | ···················· | ·A·················· | ···················· | ···················· | ···················· | ···················· | |
| G | 423 hCoV-19/USA/MN-MDH-1547/2020\|EPI_ISL_527612\|2020-0 | ···················· | ·A·················· | ···················· | ···················· | ···················· | ···················· | |
| G | 424 hCoV-19/USA/MN-MDH-1548/2020\|EPI_ISL_527613\|2020-0 | ···················· | ·A·················· | ···················· | ···················· | ···················· | ···················· | |
| G | 425 hCoV-19/USA/MN-MDH-1549/2020\|EPI_ISL_527614\|2020-0 | ···················· | ·A·················· | ···················· | ···················· | ···················· | ···················· | |
| G | 426 hCoV-19/USA/MN-MDH-1550/2020\|EPI_ISL_527615\|2020-0 | ···················· | ·A·················· | ···················· | ···················· | ···················· | ···················· | |
| G | 427 hCoV-19/USA/MN-MDH-1551/2020\|EPI_ISL_527616\|2020-0 | ···················· | ·A·················· | ···················· | ···················· | ···················· | ···················· | |
| G | 428 hCoV-19/USA/MN-MDH-1552/2020\|EPI_ISL_527617\|2020-0 | ···················· | ·A·················· | ···················· | ···················· | ···················· | ···················· | |
| G | 429 hCoV-19/USA/MN-MDH-1553/2020\|EPI_ISL_527618\|2020-0 | ···················· | ·A·················· | ···················· | ···················· | ···················· | ···················· | |
| G | 430 hCoV-19/USA/MN-MDH-1554/2020\|EPI_ISL_527619\|2020-0 | ···················· | ·A·················· | ···················· | ···················· | ···················· | ···················· | |
| G | 431 hCoV-19/USA/MN-MDH-1555/2020\|EPI_ISL_527620\|2020-0 | ···················· | ·A·················· | ···················· | ···················· | ···················· | ···················· | |
| G | 432 hCoV-19/USA/MN-MDH-1556/2020\|EPI_ISL_527621\|2020-0 | ···················· | ·A·················· | ···················· | ···················· | ···················· | ···················· | |
| G | 433 hCoV-19/USA/MN-MDH-1557/2020\|EPI_ISL_527622\|2020-0 | ···················· | ···················· | ···················· | ···················· | ···················· | ···················· | |
| G | 434 hCoV-19/USA/MN-MDH-1558/2020\|EPI_ISL_527623\|2020-0 | ···················· | ···················· | ···················· | ···················· | ···················· | ···················· | |
| G | 435 hCoV-19/USA/MN-MDH-1559/2020\|EPI_ISL_527624\|2020-0 | ···················· | ···················· | ···················· | ···················· | ···················· | ···················· | |

| | | | | | | | | |
|---|---|---|---|---|---|---|---|---|
| G | 437 hCoV-19/USA/MN-MDH-1561/2020\|EPI_ISL_527626\|2020-0 | ···················· | ·A·················· | ···················· | ···················· | ···················· | ···················· | |
| G | 438 hCoV-19/USA/MN-MDH-1562/2020\|EPI_ISL_527627\|2020-0 | ···················· | ···················· | ···················· | ···················· | NNN················· | ···················· | 1 |
| G | 439 hCoV-19/USA/MN-MDH-1563/2020\|EPI_ISL_527628\|2020-0 | ···················· | ·A·················· | ···················· | ···················· | ···················· | ···················· | |
| G | 440 hCoV-19/USA/MN-MDH-1564/2020\|EPI_ISL_527629\|2020-0 | ···················· | ·A·················· | ···················· | ···················· | ·NN················· | ···················· | |
| G | 441 hCoV-19/USA/AR-CDC-4206/2020\|EPI_ISL_527633\|2020-0 | ···················· | ···················· | ···················· | ···················· | ···················· | ···················· | |
| G | 442 hCoV-19/USA/AR-CDC-4212/2020\|EPI_ISL_527634\|2020-0 | ···················· | ···················· | ···················· | ···················· | ···················· | ···················· | |
| G | 443 hCoV-19/USA/GA-CDC-6420/2020\|EPI_ISL_527660\|2020-0 | ···················· | ···················· | ···················· | ···················· | ···················· | ···················· | |
| G | 444 hCoV-19/USA/GA-CDC-6424/2020\|EPI_ISL_527661\|2020-0 | ···················· | ···················· | ···················· | ···················· | ···················· | ···················· | |
| G | 445 hCoV-19/USA/GA-CDC-6427/2020\|EPI_ISL_527662\|2020-0 | ···················· | ·A·················· | ···················· | ···················· | ···················· | ···················· | |
| G | 446 hCoV-19/USA/GA-CDC-6431/2020\|EPI_ISL_527663\|2020-0 | ···················· | ···················· | ···················· | ···················· | ···················· | ···················· | |
| G | 447 hCoV-19/USA/MN-CDC-6192/2020\|EPI_ISL_527669\|2020-0 | ···················· | ···················· | ···················· | ···················· | ···················· | ···················· | |
| G | 448 hCoV-19/USA/MN-CDC-6195/2020\|EPI_ISL_527670\|2020-0 | ···················· | ···················· | ···················· | ···················· | ···················· | ···················· | |
| G | 449 hCoV-19/USA/MN-CDC-6201/2020\|EPI_ISL_527671\|2020-0 | ···················· | ···················· | ···················· | ···················· | ···················· | ···················· | |
| G | 450 hCoV-19/USA/MN-CDC-6205/2020\|EPI_ISL_527672\|2020-0 | ···················· | ···················· | ···················· | ···················· | ···················· | ···················· | |
| G | 451 hCoV-19/USA/MN-CDC-6208/2020\|EPI_ISL_527673\|2020-0 | ···················· | ···················· | ···················· | ···················· | ···················· | ···················· | |
| G | 452 hCoV-19/USA/MN-CDC-6211/2020\|EPI_ISL_527674\|2020-0 | ···················· | ···················· | ···················· | ···················· | ···················· | ···················· | |
| G | 453 hCoV-19/USA/MN-CDC-6214/2020\|EPI_ISL_527675\|2020-0 | ···················· | ···················· | ···················· | ···················· | ···················· | ···················· | |
| G | 454 hCoV-19/USA/MN-CDC-6215/2020\|EPI_ISL_527676\|2020-0 | ···················· | ···················· | ···················· | ···················· | ···················· | ···················· | |
| G | 455 hCoV-19/USA/MN-CDC-6218/2020\|EPI_ISL_527677\|2020-0 | ···················· | ···················· | ···················· | ···················· | ···················· | ···················· | |
| G | 456 hCoV-19/USA/MN-CDC-6221/2020\|EPI_ISL_527678\|2020-0 | ···················· | ···················· | ···················· | ···················· | ···················· | ···················· | |
| G | 457 hCoV-19/USA/MN-CDC-6227/2020\|EPI_ISL_527679\|2020-0 | ···················· | ···················· | ···················· | ···················· | ···················· | ···················· | |
| G | 458 hCoV-19/USA/MN-CDC-6233/2020\|EPI_ISL_527680\|2020-0 | ···················· | ···················· | ···················· | ···················· | ···················· | ···················· | |
| G | 459 hCoV-19/USA/MN-CDC-6237/2020\|EPI_ISL_527681\|2020-0 | ···················· | ···················· | ···················· | ···················· | ···················· | ···················· | |
| G | 460 hCoV-19/USA/MN-CDC-6244/2020\|EPI_ISL_527682\|2020-0 | ···················· | ···················· | ···················· | ···················· | ···················· | ···················· | |
| G | 461 hCoV-19/USA/MN-CDC-6247/2020\|EPI_ISL_527683\|2020-0 | ···················· | ···················· | ···················· | ···················· | ···················· | ···················· | |
| G | 462 hCoV-19/USA/MN-CDC-6250/2020\|EPI_ISL_527684\|2020-0 | ···················· | ···················· | ···················· | ···················· | ···················· | ···················· | |
| G | 463 hCoV-19/USA/MN-CDC-6256/2020\|EPI_ISL_527685\|2020-0 | ···················· | ···················· | ···················· | ···················· | ···················· | ···················· | |
| G | 464 hCoV-19/USA/MN-CDC-6259/2020\|EPI_ISL_527686\|2020-0 | ···················· | ···················· | ···················· | ···················· | ···················· | ···················· | |
| G | 465 hCoV-19/USA/MN-CDC-6262/2020\|EPI_ISL_527687\|2020-0 | ···················· | ···················· | ···················· | ···················· | ···················· | ···················· | |
| G | 466 hCoV-19/USA/MN-CDC-6265/2020\|EPI_ISL_527688\|2020-0 | ···················· | ···················· | ···················· | ···················· | ···················· | ···················· | |
| G | 467 hCoV-19/USA/MN-CDC-6268/2020\|EPI_ISL_527689\|2020-0 | ···················· | ···················· | ···················· | ···················· | ···················· | ···················· | |
| G | 468 hCoV-19/USA/MN-CDC-6274/2020\|EPI_ISL_527690\|2020-0 | ···················· | ···················· | ···················· | ···················· | ···················· | ···················· | |
| G | 469 hCoV-19/USA/MN-CDC-6277/2020\|EPI_ISL_527691\|2020-0 | ···················· | ···················· | ···················· | ···················· | ···················· | ···················· | |
| G | 470 hCoV-19/USA/MN-CDC-6281/2020\|EPI_ISL_527692\|2020-0 | ···················· | ···················· | ···················· | ···················· | ···················· | ···················· | |
| G | 471 hCoV-19/USA/MN-CDC-6286/2020\|EPI_ISL_527693\|2020-0 | ···················· | ···················· | ···················· | ···················· | ···················· | ···················· | |
| G | 472 hCoV-19/USA/MN-CDC-6290/2020\|EPI_ISL_527694\|2020-0 | ···················· | ···················· | ···················· | ···················· | ···················· | ···················· | |
| G | 473 hCoV-19/USA/MN-CDC-6295/2020\|EPI_ISL_527695\|2020-0 | ···················· | ···················· | ···················· | ···················· | ···················· | ···················· | |
| G | 474 hCoV-19/USA/MN-CDC-6298/2020\|EPI_ISL_527696\|2020-0 | ···················· | ···················· | ···················· | ···················· | ···················· | ···················· | |
| G | 475 hCoV-19/USA/MN-CDC-6302/2020\|EPI_ISL_527697\|2020-0 | ···················· | ···················· | ···················· | ···················· | ···················· | ···················· | |
| G | 476 hCoV-19/USA/MN-CDC-6305/2020\|EPI_ISL_527698\|2020-0 | ···················· | ···················· | ···················· | ···················· | ···················· | ···················· | |
| G | 477 hCoV-19/USA/MN-CDC-6312/2020\|EPI_ISL_527699\|2020-0 | ···················· | ···················· | ···················· | ···················· | ···················· | ···················· | |
| G | 478 hCoV-19/Italy/ABR-IZSGC-TE4884/2020\|EPI_ISL_528919 | ···················· | ···················· | ···················· | ···················· | ···················· | ···················· | |
| G | 479 hCoV-19/Italy/ABR-IZSGC-TE4891/2020\|EPI_ISL_528920 | ···················· | ···················· | ···················· | ···················· | ···················· | ···················· | |
| G | 480 hCoV-19/Italy/ABR-IZSGC-TE4926/2020\|EPI_ISL_528921 | ···················· | ···················· | ···················· | ···················· | ···················· | ···················· | |
| G | 481 hCoV-19/Italy/ABR-IZSGC-TE5040/2020\|EPI_ISL_528922 | ···················· | ···················· | ···················· | ···················· | ···················· | ···················· | |
| G | 482 hCoV-19/Italy/ABR-IZSGC-TE5173/2020\|EPI_ISL_528924 | ···················· | ···················· | ···················· | ···················· | ···················· | ···················· | |
| G | 483 hCoV-19/Italy/ABR-IZSGC-TE5268/2020\|EPI_ISL_528925 | ···················· | ···················· | ···················· | ···················· | ···················· | ···················· | |
| G | 484 hCoV-19/Italy/ABR-IZSGC-TE5487/2020\|EPI_ISL_528926 | ···················· | ···················· | ···················· | ···················· | ···················· | ···················· | |
| G | 485 hCoV-19/Italy/ABR-IZSGC-TE6202/2020\|EPI_ISL_528927 | ···················· | ···················· | ···················· | ···················· | ···················· | ···················· | |
| G | 486 hCoV-19/Italy/ABR-IZSGC-TE6218/2020\|EPI_ISL_528928 | ···················· | ···················· | ···················· | ···················· | ···················· | ···················· | |
| G | 487 hCoV-19/Italy/ABR-IZSGC-TE7097/2020\|EPI_ISL_528929 | ···················· | ···················· | ···················· | ···················· | ···················· | ···················· | |
| G | 488 hCoV-19/England/20202015104/2020\|EPI_ISL_528930\|20 | ···················· | ···················· | ···················· | ···················· | ···················· | ···················· | |
| G | 489 hCoV-19/Italy/LOM-IZSGC-TE30933/2020\|EPI_ISL_52893 | ···················· | ···················· | ···················· | ···················· | ···················· | ···················· | |
| G | 490 hCoV-19/Italy/LOM-IZSGC-TE31054/2020\|EPI_ISL_52893 | ···················· | ···················· | ···················· | ···················· | ···················· | ···················· | |
| G | 491 hCoV-19/Italy/LOM-IZSGC-TE31074/2020\|EPI_ISL_52893 | ···················· | ···················· | ···················· | ···················· | ···················· | ···················· | |
| G | 492 hCoV-19/Italy/LOM-IZSGC-TE31135/2020\|EPI_ISL_52893 | ···················· | ···················· | ···················· | ···················· | ···················· | ···················· | |
| G | 493 hCoV-19/Italy/LOM-IZSGC-TE31751/2020\|EPI_ISL_52893 | ···················· | ···················· | ···················· | ···················· | ···················· | ···················· | |
| G | 494 hCoV-19/Italy/LOM-IZSGC-TE31811/2020\|EPI_ISL_52893 | ···················· | ···················· | ···················· | ···················· | ···················· | ···················· | |
| G | 495 hCoV-19/Italy/LOM-IZSGC-TE31818/2020\|EPI_ISL_52894 | ···················· | ···················· | ···················· | ···················· | ···················· | ···················· | |
| G | 496 hCoV-19/Italy/LOM-IZSGC-TE35432/2020\|EPI_ISL_52894 | ···················· | ···················· | ···················· | ···················· | ···················· | ···················· | |
| G | 497 hCoV-19/Italy/LOM-IZSGC-TE35441/2020\|EPI_ISL_52894 | ···················· | ···················· | ···················· | ···················· | ···················· | ···················· | |
| G | 498 hCoV-19/Italy/LOM-IZSGC-TE35526/2020\|EPI_ISL_52894 | ···················· | ···················· | ···················· | ···················· | ···················· | ···················· | |
| G | 499 hCoV-19/Italy/LOM-IZSGC-TE35532/2020\|EPI_ISL_52894 | ···················· | ···················· | ···················· | ···················· | ···················· | ···················· | |
| G | 500 hCoV-19/Italy/LOM-IZSGC-TE35566/2020\|EPI_ISL_52894 | ···················· | ···················· | ···················· | ···················· | ···················· | ···················· | |
| G | 501 hCoV-19/Italy/LOM-IZSGC-TE35578/2020\|EPI_ISL_52894 | ···················· | ···················· | ···················· | ···················· | ···················· | ···················· | |
| G | 502 hCoV-19/Italy/LOM-IZSGC-TE38811/2020\|EPI_ISL_52894 | ···················· | ···················· | ···················· | ···················· | ···················· | ···················· | |
| G | 503 hCoV-19/Italy/LOM-IZSGC-TE38819/2020\|EPI_ISL_52894 | ···················· | ···················· | ···················· | ···················· | ···················· | ···················· | |
| G | 504 hCoV-19/Italy/LOM-IZSGC-TE38845/2020\|EPI_ISL_52894 | ···················· | ···················· | ···················· | ···················· | ···················· | ···················· | |
| G | 505 hCoV-19/England/20166019404/2020\|EPI_ISL_528950\|20 | ···················· | ···················· | ···················· | ···················· | ···················· | ···················· | |
| G | 506 hCoV-19/Italy/ABR-IZSGC-TE10002/2020\|EPI_ISL_52899 | ···················· | ···················· | ···················· | ···················· | ···················· | ···················· | |
| G | 507 hCoV-19/Italy/ABR-IZSGC-TE10007/2020\|EPI_ISL_52899 | ···················· | ···················· | ···················· | ···················· | ···················· | ···················· | |
| G | 508 hCoV-19/Italy/ABR-IZSGC-TE34742/2020\|EPI_ISL_52899 | ···················· | ···················· | ···················· | ···················· | ···················· | ···················· | |
| G | 509 hCoV-19/Italy/ABR-IZSGC-TE46129/2020\|EPI_ISL_52899 | ···················· | ···················· | ···················· | ···················· | ···················· | ···················· | |
| G | 510 hCoV-19/USA/WA-RML-1/2020\|EPI_ISL_641314\|2020-04-2 | ···················· | ···················· | ···················· | ···················· | ···················· | ···················· | |
| G | 511 hCoV-19/USA/WA-RML-3/2020\|EPI_ISL_641316\|2020-05-2 | ···················· | ···················· | ···················· | ···················· | ···················· | ···················· | |
| G | 512 hCoV-19/USA/WA-RML-4/2020\|EPI_ISL_641317\|2020-06-1 | ···················· | ···················· | ···················· | ···················· | ···················· | ···················· | |
| G | 513 hCoV-19/England/CAMB-1B5B17/2020\|EPI_ISL_516993\|20 | ···················· | ···················· | ···················· | ···················· | ···················· | ···················· | |
| G | 514 hCoV-19/England/CAMB-1B5B35/2020\|EPI_ISL_516995\|20 | ···················· | ···················· | ···················· | ···················· | ···················· | ···················· | |
| G | 515 hCoV-19/England/CAMB-1B5B44/2020\|EPI_ISL_516996\|20 | ···················· | ···················· | ···················· | ···················· | ···················· | ···················· | |
| G | 516 hCoV-19/England/CAMB-1B5B71/2020\|EPI_ISL_516999\|20 | ···················· | ···················· | ···················· | ···················· | ···················· | ···················· | |

G 517 hCoV-19/Australia/VIC4649/2020|EPI_ISL_519040|2020 .................... .................... .................... .................... .................... ....................
G 518 hCoV-19/Australia/VIC4650/2020|EPI_ISL_519041|2020 .................... .................... .................... .................... .................... ....................
G 519 hCoV-19/Australia/VIC4651/2020|EPI_ISL_519042|2020 .................... .................... .................... .................... .................... ....................
G 520 hCoV-19/Australia/VIC4661/2020|EPI_ISL_519050|2020 .................... .................... .................... .................... .................... ....................
G 521 hCoV-19/Australia/VIC4662/2020|EPI_ISL_519051|2020 .................... .................... .................... .................... .................... ....................
G 522 hCoV-19/Australia/VIC4663/2020|EPI_ISL_519052|2020 .................... .................... .................... .................... .................... ....................
G 523 hCoV-19/Australia/VIC4665/2020|EPI_ISL_519053|2020 .................... .................... .................... .................... .................... ....................
G 524 hCoV-19/Australia/VIC4666/2020|EPI_ISL_519054|2020 .................... .................... .................... .................... .................... ....................
G 525 hCoV-19/Australia/VIC4667/2020|EPI_ISL_519055|2020 .................... .................... .................... .................... .................... ....................
G 526 hCoV-19/Australia/VIC4668/2020|EPI_ISL_519056|2020 .................... .................... .................... .................... .................... ....................
G 527 hCoV-19/Australia/VIC4669/2020|EPI_ISL_519057|2020 .................... .................... .................... .................... .................... ....................
G 528 hCoV-19/Australia/VIC4670/2020|EPI_ISL_519058|2020 .................... .................... .................... .................... .................... ....................
G 529 hCoV-19/Australia/VIC4671/2020|EPI_ISL_519059|2020 .................... .................... .................... .................... .................... ....................
G 530 hCoV-19/Australia/VIC4815/2020|EPI_ISL_519200|2020 .................... .................... .................... .................... .................... ....................
G 531 hCoV-19/Australia/VIC4816/2020|EPI_ISL_519201|2020 .................... .................... .................... .................... .................... ....................
G 532 hCoV-19/Australia/VIC4818/2020|EPI_ISL_519203|2020 .................... .................... .................... .................... .................... ....................
G 533 hCoV-19/Australia/VIC4819/2020|EPI_ISL_519204|2020 .................... .................... .................... .................... .................... ....................
G 534 hCoV-19/Australia/VIC4820/2020|EPI_ISL_519205|2020 .................... .................... .................... .................... .................... ....................
G 535 hCoV-19/Australia/VIC4821/2020|EPI_ISL_519206|2020 .................... .................... .................... .................... .................... ....................
G 536 hCoV-19/Australia/VIC4822/2020|EPI_ISL_519207|2020 .................... .................... .................... .................... .................... ....................
G 537 hCoV-19/Australia/VIC4823/2020|EPI_ISL_519208|2020 .................... .................... .................... .................... .................... ....................
G 538 hCoV-19/Australia/VIC4824/2020|EPI_ISL_519209|2020 .................... .................... .................... .................... .................... ....................
G 539 hCoV-19/Australia/VIC4826/2020|EPI_ISL_519210|2020 .................... .................... .................... .................... .................... ....................
G 540 hCoV-19/Australia/VIC4827/2020|EPI_ISL_519211|2020 .................... .................... .................... .................... .................... ....................
G 541 hCoV-19/Australia/VIC4828/2020|EPI_ISL_519212|2020 .................... .................... .................... .................... .................... ....................
G 542 hCoV-19/Australia/VIC4829/2020|EPI_ISL_519213|2020 .................... .................... .................... .................... .................... ....................
G 543 hCoV-19/Australia/VIC4830/2020|EPI_ISL_519214|2020 .................... .................... .................... .................... .................... ....................
G 544 hCoV-19/Australia/VIC4831/2020|EPI_ISL_519215|2020 .................... .................... .................... .................... .................... ....................
G 545 hCoV-19/Australia/VIC4832/2020|EPI_ISL_519216|2020 .................... .................... .................... .................... .................... ....................
G 546 hCoV-19/Australia/VIC4833/2020|EPI_ISL_519217|2020 .................... .................... .................... .................... .................... ....................
G 547 hCoV-19/Australia/VIC4835/2020|EPI_ISL_519219|2020 .................... .................... .................... .................... .................... ....................
G 548 hCoV-19/Australia/VIC5262/2020|EPI_ISL_520027|2020 .................... .................... .................... .................... .................... ....................
G 549 hCoV-19/Australia/VIC5263/2020|EPI_ISL_520028|2020 .................... .................... .................... .................... .................... ....................
G 550 hCoV-19/Australia/VIC5264/2020|EPI_ISL_520029|2020 .................... .................... .................... .................... .................... ....................
G 551 hCoV-19/Australia/VIC5265/2020|EPI_ISL_520030|2020 .................... .................... .................... .................... .................... ....................
G 552 hCoV-19/Australia/VIC5266/2020|EPI_ISL_520031|2020 .................... .................... .................... .................... .................... ....................
G 553 hCoV-19/Australia/VIC5267/2020|EPI_ISL_520032|2020 .................... .................... .................... .................... .................... ....................
G 554 hCoV-19/Australia/VIC5268/2020|EPI_ISL_520033|2020 .................... .................... .................... .................... .................... ....................
G 555 hCoV-19/Australia/VIC5269/2020|EPI_ISL_520034|2020 .................... .................... .................... .................... .................... ....................
G 556 hCoV-19/Australia/VIC5270/2020|EPI_ISL_520035|2020 .................... .................... .................... .................... .................... ....................
G 557 hCoV-19/Australia/VIC5271/2020|EPI_ISL_520036|2020 .................... .................... .................... .................... .................... ....................
G 558 hCoV-19/Australia/VIC5272/2020|EPI_ISL_520037|2020 .................... .................... .................... .................... .................... ....................
G 559 hCoV-19/Australia/VIC5273/2020|EPI_ISL_520038|2020 .................... .................... .................... .................... .................... ....................
G 560 hCoV-19/Australia/VIC5274/2020|EPI_ISL_520039|2020 .................... .................... .................... .................... .................... ....................
G 561 hCoV-19/Australia/VIC5275/2020|EPI_ISL_520040|2020 .................... .................... .................... .................... .................... ....................
G 562 hCoV-19/Australia/VIC5276/2020|EPI_ISL_520041|2020 .................... .................... .................... .................... .................... ....................
G 563 hCoV-19/Australia/VIC5277/2020|EPI_ISL_520042|2020 .................... .................... .................... .................... .................... ....................
G 564 hCoV-19/Australia/VIC5278/2020|EPI_ISL_520043|2020 .................... .................... .................... .................... .................... ....................
G 565 hCoV-19/Australia/VIC5279/2020|EPI_ISL_520044|2020 .................... .................... .................... .................... .................... ....................
G 566 hCoV-19/Australia/VIC5280/2020|EPI_ISL_520045|2020 .................... .................... .................... .................... .................... ....................
G 567 hCoV-19/Australia/VIC5281/2020|EPI_ISL_520046|2020 .................... .................... .................... .................... .................... ....................
G 568 hCoV-19/Australia/VIC5282/2020|EPI_ISL_520047|2020 .................... .................... .................... .................... .................... ....................
G 569 hCoV-19/Australia/VIC5283/2020|EPI_ISL_520048|2020 .................... .................... .................... .................... .................... ....................
G 570 hCoV-19/Australia/VIC5449/2020|EPI_ISL_520200|2020 .................... .................... .................... .................... .................... ....................
G 571 hCoV-19/Australia/VIC5452/2020|EPI_ISL_520202|2020 .................... .................... .................... .................... .................... ....................
G 572 hCoV-19/Australia/VIC6596/2020|EPI_ISL_521358|2020 .................... .................... .................... .................... .................... ....................
G 573 hCoV-19/Australia/VIC6597/2020|EPI_ISL_521359|2020 .................... .................... .................... .................... .................... ....................
G 574 hCoV-19/Australia/VIC6598/2020|EPI_ISL_521360|2020 .................... .................... .................... .................... .................... ....................
G 575 hCoV-19/Australia/VIC6599/2020|EPI_ISL_521361|2020 .................... .................... .................... .................... .................... ....................
G 576 hCoV-19/Australia/VIC6600/2020|EPI_ISL_521362|2020 .................... .................... .................... .................... .................... ....................
G 577 hCoV-19/Australia/VIC6601/2020|EPI_ISL_521363|2020 .................... .................... .................... .................... .................... ....................
G 578 hCoV-19/Australia/VIC6602/2020|EPI_ISL_521364|2020 .................... .................... .................... .................... .................... ....................
G 579 hCoV-19/Australia/VIC6603/2020|EPI_ISL_521365|2020 .................... .................... .................... .................... .................... ....................
G 580 hCoV-19/Australia/VIC6604/2020|EPI_ISL_521366|2020 .................... .................... .................... .................... .................... ....................
G 581 hCoV-19/Australia/VIC6605/2020|EPI_ISL_521367|2020 .................... .................... .................... .................... .................... ....................
G 582 hCoV-19/Australia/VIC6606/2020|EPI_ISL_521368|2020 .................... .................... .................... .................... .................... ....................
G 583 hCoV-19/Australia/VIC6607/2020|EPI_ISL_521369|2020 .................... .................... .................... .................... .................... ....................
G 584 hCoV-19/Australia/VIC6608/2020|EPI_ISL_521370|2020 .................... .................... .................... .................... .................... ....................
G 585 hCoV-19/Australia/VIC6609/2020|EPI_ISL_521371|2020 .................... .................... .................... .................... .................... ....................
G 586 hCoV-19/Australia/VIC6610/2020|EPI_ISL_521372|2020 .................... .................... .................... .................... .................... ....................
G 587 hCoV-19/Australia/VIC6612/2020|EPI_ISL_521374|2020 .................... .................... .................... .................... .................... ....................
G 588 hCoV-19/Australia/VIC6614/2020|EPI_ISL_521376|2020 .................... .................... .................... .................... .................... ....................
G 589 hCoV-19/Australia/VIC6615/2020|EPI_ISL_521377|2020 .................... .................... .................... .................... .................... ....................
G 590 hCoV-19/Australia/VIC6617/2020|EPI_ISL_521379|2020 .................... .................... .................... .................... .................... ....................
G 591 hCoV-19/Australia/VIC2654/2020|EPI_ISL_521500|2020 .................... .................... .................... .................... .................... ....................
G 592 hCoV-19/Australia/VIC2656/2020|EPI_ISL_521502|2020 .................... .................... .................... .................... .................... ....................
G 593 hCoV-19/Australia/VIC2657/2020|EPI_ISL_521503|2020 .................... .................... .................... .................... .................... ....................
G 594 hCoV-19/Australia/VIC2658/2020|EPI_ISL_521504|2020 .................... .................... .................... .................... .................... ....................
G 595 hCoV-19/Australia/VIC2659/2020|EPI_ISL_521505|2020 .................... .................... .................... .................... .................... ....................
G 596 hCoV-19/Australia/VIC2660/2020|EPI_ISL_521506|2020 .................... .................... .................... .................... .................... ....................

```
G  597 hCoV-19/Australia/VIC2661/2020|EPI_ISL_521507|2020   ····················  ····················  ·····················  ····················  ····················  ····················
G  598 hCoV-19/Australia/VIC2664/2020|EPI_ISL_521508|2020   ····················  ····················  ·····················  ····················  ····················  ····················
G  599 hCoV-19/Australia/VIC2667/2020|EPI_ISL_521509|2020   ····················  ····················  ·····················  ····················  ····················  ····················
G  600 hCoV-19/Australia/VIC2668/2020|EPI_ISL_521510|2020   ····················  ····················  ·····················  ····················  ····················  ····················
G  601 hCoV-19/Australia/VIC2669/2020|EPI_ISL_521511|2020   ····················  ····················  ·····················  ····················  ····················  ····················
G  602 hCoV-19/Australia/VIC2670/2020|EPI_ISL_521512|2020   ····················  ····················  ·····················  ····················  ····················  ····················
G  603 hCoV-19/Australia/VIC2672/2020|EPI_ISL_521514|2020   ····················  ····················  ·····················  ····················  ····················  ····················
G  604 hCoV-19/Australia/VIC2673/2020|EPI_ISL_521515|2020   ····················  ····················  ·····················  ····················  ····················  ····················
G  605 hCoV-19/Australia/VIC2674/2020|EPI_ISL_521516|2020   ····················  ····················  ·····················  ····················  ····················  ····················
G  606 hCoV-19/Australia/VIC2676/2020|EPI_ISL_521517|2020   ····················  ····················  ·····················  ····················  ····················  ····················
G  607 hCoV-19/Australia/VIC2677/2020|EPI_ISL_521518|2020   ····················  ····················  ·····················  ····················  ····················  ····················
G  608 hCoV-19/Australia/VIC2678/2020|EPI_ISL_521519|2020   ····················  ····················  ·····················  ····················  ····················  ····················
G  609 hCoV-19/Australia/VIC2679/2020|EPI_ISL_521520|2020   ····················  ····················  ·····················  ····················  ····················  ····················
G  610 hCoV-19/Australia/VIC2680/2020|EPI_ISL_521521|2020   ····················  ····················  ·····················  ····················  ····················  ····················
G  611 hCoV-19/Australia/VIC2681/2020|EPI_ISL_521522|2020   ····················  ····················  ·····················  ····················  ····················  ····················
G  612 hCoV-19/Australia/VIC2682/2020|EPI_ISL_521523|2020   ····················  ····················  ·····················  ····················  ····················  ····················
G  613 hCoV-19/Australia/VIC2683/2020|EPI_ISL_521524|2020   ····················  ····················  ·····················  ····················  ····················  ····················
G  614 hCoV-19/Australia/VIC2684/2020|EPI_ISL_521525|2020   ····················  ····················  ·····················  ····················  ····················  ····················
G  615 hCoV-19/Australia/VIC2685/2020|EPI_ISL_521526|2020   ····················  ····················  ·····················  ····················  ····················  ····················
G  616 hCoV-19/Australia/VIC2686/2020|EPI_ISL_521527|2020   ····················  ····················  ·····················  ····················  ····················  ····················
G  617 hCoV-19/Australia/VIC2687/2020|EPI_ISL_521528|2020   ····················  ····················  ·····················  ····················  ····················  ····················
G  618 hCoV-19/Australia/VIC2689/2020|EPI_ISL_521529|2020   ····················  ····················  ·····················  ····················  ····················  ····················
G  619 hCoV-19/Australia/VIC2690/2020|EPI_ISL_521530|2020   ····················  ····················  ·····················  ····················  ····················  ····················
G  620 hCoV-19/Australia/VIC2691/2020|EPI_ISL_521531|2020   ····················  ····················  ·····················  ····················  ····················  ····················
G  621 hCoV-19/Australia/VIC2692/2020|EPI_ISL_521532|2020   ····················  ····················  ·····················  ····················  ····················  ····················
G  622 hCoV-19/Australia/VIC2693/2020|EPI_ISL_521533|2020   ····················  ····················  ·····················  ····················  ····················  ····················
G  625 hCoV-19/Australia/TAS263/2020|EPI_ISL_522691|2020-   ····················  ····················  ·····················  ····················  ····················  ····················
G  630 hCoV-19/Australia/TAS268/2020|EPI_ISL_522696|2020-   ····················  ····················  ·····················  ····················  ····················  ····················
G  631 hCoV-19/Australia/TAS269/2020|EPI_ISL_522697|2020-   ····················  ····················  ·····················  ····················  ····················  ····················
G  633 hCoV-19/USA/VA-DCLS-0935/2020|EPI_ISL_522800|2020-   ····················  ····················  ·····················  ····················  ····················  ····················
G  634 hCoV-19/USA/VA-DCLS-0936/2020|EPI_ISL_522801|2020-   ····················  ····················  ·····················  ····················  ····················  ····················
G  635 hCoV-19/USA/VA-DCLS-0938/2020|EPI_ISL_522802|2020-   ····················  ····················  ·····················  ····················  ····················  ····················
G  636 hCoV-19/USA/VA-DCLS-0942/2020|EPI_ISL_522803|2020-   ····················  ····················  ·····················  ····················  ····················  ····················
G  637 hCoV-19/USA/VA-DCLS-0944/2020|EPI_ISL_522804|2020-   ····················  ····················  ·····················  ····················  ····················  ····················
G  638 hCoV-19/USA/VA-DCLS-0948/2020|EPI_ISL_522805|2020-   ····················  ····················  ·····················  ····················  ····················  ····················
G  639 hCoV-19/USA/VA-DCLS-0795/2020|EPI_ISL_522806|2020-   ····················  ····················  ·····················  ····················  ····················  ····················
G  640 hCoV-19/USA/VA-DCLS-0796/2020|EPI_ISL_522807|2020-   ····················  ····················  ·····················  ····················  ····················  ····················
G  641 hCoV-19/USA/VA-DCLS-0797/2020|EPI_ISL_522808|2020-   ····················  ····················  ·····················  ····················  ····················  ····················
G  642 hCoV-19/USA/VA-DCLS-0798/2020|EPI_ISL_522809|2020-   ····················  ····················  ·····················  ····················  ····················  ····················
G  643 hCoV-19/USA/VA-DCLS-0799/2020|EPI_ISL_522810|2020-   ····················  ····················  ·····················  ····················  ····················  ····················
G  644 hCoV-19/USA/VA-DCLS-0801/2020|EPI_ISL_522811|2020-   ····················  ····················  ·····················  ····················  ····················  ····················
G  645 hCoV-19/USA/VA-DCLS-0803/2020|EPI_ISL_522812|2020-   ····················  ····················  ·····················  ····················  ····················  ····················
G  646 hCoV-19/USA/VA-DCLS-0804/2020|EPI_ISL_522813|2020-   ····················  ····················  ·····················  ····················  ····················  ····················
G  647 hCoV-19/USA/VA-DCLS-0805/2020|EPI_ISL_522814|2020-   ····················  ····················  ·····················  ····················  ····················  ····················
G  648 hCoV-19/USA/VA-DCLS-0807/2020|EPI_ISL_522815|2020-   ····················  ····················  ·····················  ····················  ····················  ····················
G  649 hCoV-19/USA/VA-DCLS-0808/2020|EPI_ISL_522816|2020-   ····················  ····················  ·····················  ····················  ····················  ····················
G  650 hCoV-19/USA/VA-DCLS-0810/2020|EPI_ISL_522817|2020-   ····················  ····················  ·····················  ····················  ····················  ····················
G  651 hCoV-19/USA/VA-DCLS-0811/2020|EPI_ISL_522818|2020-   ····················  ····················  ·····················  ····················  ····················  ····················
G  652 hCoV-19/USA/VA-DCLS-0812/2020|EPI_ISL_522819|2020-   ····················  ····················  ·····················  ····················  ····················  ····················
G  653 hCoV-19/USA/VA-DCLS-1153/2020|EPI_ISL_522820|2020-   ····················  ····················  ·····················  ····················  ····················  ····················
G  654 hCoV-19/USA/VA-DCLS-1154/2020|EPI_ISL_522821|2020-   ····················  ····················  ·····················  ····················  ····················  ····················
G  655 hCoV-19/USA/VA-DCLS-1155/2020|EPI_ISL_522822|2020-   ····················  ····················  ·····················  ····················  ····················  ····················
G  656 hCoV-19/USA/VA-DCLS-1156/2020|EPI_ISL_522823|2020-   ····················  ····················  ·····················  ····················  ····················  ····················
G  657 hCoV-19/USA/MD-MDH-0088/2020|EPI_ISL_522824|2020-0   ····················  ····················  ·····················  ····················  ····················  ····················
G  658 hCoV-19/USA/MD-MDH-0089/2020|EPI_ISL_522825|2020-0   ····················  ····················  ·····················  ····················  ····················  ····················
G  659 hCoV-19/USA/MD-MDH-0090/2020|EPI_ISL_522826|2020-0   ····················  ····················  ·····················  ····················  ····················  ····················
G  660 hCoV-19/USA/MD-MDH-0091/2020|EPI_ISL_522827|2020-0   ····················  ····················  ·····················  ····················  ····················  ····················
G  661 hCoV-19/USA/MD-MDH-0092/2020|EPI_ISL_522828|2020-0   ····················  ····················  ·····················  ····················  ····················  ····················
G  662 hCoV-19/USA/MD-MDH-0093/2020|EPI_ISL_522829|2020-0   ····················  ····················  ·····················  ····················  ····················  ····················
G  663 hCoV-19/USA/MD-MDH-0094/2020|EPI_ISL_522830|2020-0   ····················  ····················  ·····················  ····················  ····················  ····················
G  664 hCoV-19/USA/MD-MDH-0095/2020|EPI_ISL_522831|2020-0   ····················  ····················  ·····················  ····················  ····················  ····················
G  665 hCoV-19/USA/MD-MDH-0096/2020|EPI_ISL_522832|2020-0   ····················  ····················  ·····················  ····················  ····················  ····················
G  666 hCoV-19/USA/MD-MDH-0098/2020|EPI_ISL_522833|2020-0   ····················  ····················  ·····················  ····················  ····················  ····················
G  667 hCoV-19/USA/MD-MDH-0099/2020|EPI_ISL_522834|2020-0   ····················  ····················  ·····················  ····················  ····················  ····················
G  668 hCoV-19/USA/MD-MDH-0101/2020|EPI_ISL_522836|2020-0   ····················  ····················  ·····················  ····················  ····················  ····················
G  669 hCoV-19/USA/MD-MDH-0102/2020|EPI_ISL_522837|2020-0   ····················  ····················  ·····················  ····················  ····················  ····················
G  670 hCoV-19/USA/MD-MDH-0103/2020|EPI_ISL_522838|2020-0   ····················  ····················  ·····················  ····················  ····················  ····················
G  671 hCoV-19/USA/MD-MDH-0104/2020|EPI_ISL_522839|2020-0   ····················  ····················  ·····················  ····················  ····················  ····················
G  672 hCoV-19/USA/MD-MDH-0105/2020|EPI_ISL_522840|2020-0   ····················  ····················  ·····················  ····················  ····················  ····················
G  673 hCoV-19/USA/MD-MDH-0107/2020|EPI_ISL_522842|2020-0   ····················  ····················  ·····················  ····················  ····················  ····················
G  674 hCoV-19/USA/MD-MDH-0108/2020|EPI_ISL_522843|2020-0   ····················  ····················  ·····················  ····················  ····················  ····················
G  675 hCoV-19/USA/MD-MDH-0109/2020|EPI_ISL_522844|2020-0   ····················  ····················  ·····················  ····················  ····················  ····················
G  676 hCoV-19/USA/MD-MDH-0110/2020|EPI_ISL_522845|2020-0   ····················  ····················  ·····················  ····················  ····················  ····················
G  677 hCoV-19/USA/MD-MDH-0112/2020|EPI_ISL_522846|2020-0   ····················  ····················  ·····················  ····················  ····················  ····················
G  678 hCoV-19/USA/MD-MDH-0115/2020|EPI_ISL_522848|2020-0   ····················  ····················  ·····················  ····················  ····················  ····················
G  679 hCoV-19/USA/MD-MDH-0116/2020|EPI_ISL_522849|2020-0   ····················  ····················  ·····················  ····················  ····················  ····················
G  680 hCoV-19/USA/MD-MDH-0117/2020|EPI_ISL_522850|2020-0   ····················  ····················  ·····················  ····················  ····················  ····················
G  681 hCoV-19/USA/MD-MDH-0118/2020|EPI_ISL_522851|2020-0   ····················  ····················  ·····················  ····················  ····················  ····················
G  682 hCoV-19/USA/MD-MDH-0119/2020|EPI_ISL_522852|2020-0   ····················  ····················  ·····················  ····················  ····················  ····················
G  683 hCoV-19/USA/MD-MDH-0120/2020|EPI_ISL_522853|2020-0   ····················  ····················  ·····················  ····················  ····················  ····················
```

| | | | | | | | |
|---|---|---|---|---|---|---|---|
| G | 684 hCoV-19/USA/MD-MDH-0121/2020\|EPI_ISL_522854\|2020-0 | ···················· | ···················· | ····················· | ···················· | ···················· | ···················· |
| G | 685 hCoV-19/Italy/VEN-IZSVe-101-12/2020\|EPI_ISL_522855 | ···················· | ···················· | ····················· | ···················· | ···················· | ···················· |
| G | 686 hCoV-19/Italy/VEN-IZSVe-46-41/2020\|EPI_ISL_522856\| | ···················· | ···················· | ····················· | ···················· | ···················· | ···················· |
| G | 687 hCoV-19/Italy/VEN-IZSVe-50893/2020\|EPI_ISL_522857\| | ···················· | ···················· | ····················· | ···················· | ···················· | ···················· |
| G | 688 hCoV-19/Italy/VEN-IZSVe-51-5/2020\|EPI_ISL_522858\|2 | ···················· | ···················· | ····················· | ···················· | ···················· | ···················· |
| G | 689 hCoV-19/Italy/VEN-IZSVe-51635/2020\|EPI_ISL_522859\| | ···················· | ···················· | ····················· | ···················· | ···················· | ···················· |
| G | 690 hCoV-19/Italy/VEN-IZSVe-54-17/2020\|EPI_ISL_522860\| | ···················· | ···················· | ····················· | ···················· | ···················· | ···················· |
| G | 691 hCoV-19/Italy/VEN-IZSVe-55-111/2020\|EPI_ISL_522861 | ···················· | ···················· | ····················· | ···················· | ···················· | ···················· |
| G | 692 hCoV-19/Italy/VEN-IZSVe-55-16/2020\|EPI_ISL_522862\| | ···················· | ···················· | ····················· | ···················· | ···················· | ···················· |
| G | 693 hCoV-19/Italy/VEN-IZSVe-55-18/2020\|EPI_ISL_522863\| | ···················· | ···················· | ····················· | ···················· | ···················· | ···················· |
| G | 694 hCoV-19/Italy/VEN-IZSVe-55-56/2020\|EPI_ISL_522864\| | ···················· | ···················· | ····················· | ···················· | ···················· | ···················· |
| G | 695 hCoV-19/USA/UT-UPHL-2008888/2020\|EPI_ISL_525002\|20 | ···················· | ···················· | ····················· | ···················· | ···················· | ···················· |
| G | 696 hCoV-19/USA/UT-UPHL-2009046/2020\|EPI_ISL_525060\|20 | ···················· | ···················· | ····················· | ···················· | ···················· | ···················· |
| G | 697 hCoV-19/USA/UT-UPHL-2009048/2020\|EPI_ISL_525062\|20 | ···················· | ···················· | ····················· | ···················· | ···················· | ···················· |
| G | 698 hCoV-19/USA/UT-UPHL-2009051/2020\|EPI_ISL_525065\|20 | ···················· | ···················· | ····················· | ···················· | ···················· | ···················· |
| G | 699 hCoV-19/USA/UT-UPHL-2009056/2020\|EPI_ISL_525068\|20 | ···················· | ···················· | ····················· | ···················· | ···················· | ···················· |
| G | 700 hCoV-19/USA/UT-UPHL-2009058/2020\|EPI_ISL_525070\|20 | ···················· | ···················· | ····················· | ···················· | ···················· | ···················· |
| G | 701 hCoV-19/USA/UT-UPHL-2009061/2020\|EPI_ISL_525072\|20 | ···················· | ···················· | ····················· | ···················· | ···················· | ···················· |
| G | 702 hCoV-19/USA/UT-UPHL-2009064/2020\|EPI_ISL_525074\|20 | ···················· | ···················· | ····················· | ···················· | ···················· | ···················· |
| G | 703 hCoV-19/USA/UT-UPHL-2009068/2020\|EPI_ISL_525076\|20 | ···················· | ···················· | ····················· | ···················· | ···················· | ···················· |
| G | 704 hCoV-19/USA/UT-UPHL-2009069/2020\|EPI_ISL_525077\|20 | ···················· | ···················· | ····················· | ···················· | ···················· | ···················· |
| G | 705 hCoV-19/USA/UT-UPHL-2009071/2020\|EPI_ISL_525079\|20 | ···················· | ···················· | ····················· | ···················· | ·········A·········· | ···················· |
| G | 706 hCoV-19/USA/UT-UPHL-2009072/2020\|EPI_ISL_525080\|20 | ···················· | ···················· | ····················· | ···················· | ···················· | ···················· |
| G | 707 hCoV-19/USA/UT-UPHL-2009074/2020\|EPI_ISL_525082\|20 | ···················· | ···················· | ····················· | ···················· | ···················· | ···················· |
| G | 708 hCoV-19/USA/UT-UPHL-2009075/2020\|EPI_ISL_525083\|20 | ···················· | ···················· | ····················· | ···················· | ···················· | ···················· |
| G | 709 hCoV-19/USA/UT-UPHL-2009085/2020\|EPI_ISL_525092\|20 | ···················· | ···················· | ····················· | ···················· | ···················· | ···················· |
| G | 710 hCoV-19/USA/UT-UPHL-2009088/2020\|EPI_ISL_525094\|20 | ···················· | ···················· | ····················· | ···················· | ···················· | ···················· |
| G | 711 hCoV-19/England/ALDP-97CB9C/2020\|EPI_ISL_526309\|20 | ···················· | ···················· | ····················· | ···················· | ···················· | ···················· |
| G | 712 hCoV-19/England/ALDP-97CBAB/2020\|EPI_ISL_526310\|20 | ···················· | ···················· | ····················· | ···················· | ···················· | ···················· |
| G | 713 hCoV-19/England/ALDP-97CC99/2020\|EPI_ISL_526312\|20 | ···················· | ···················· | ····················· | ···················· | ···················· | ···················· |
| G | 714 hCoV-19/England/ALDP-97CCB7/2020\|EPI_ISL_526313\|20 | ···················· | ···················· | ····················· | ···················· | ···················· | ···················· |
| G | 715 hCoV-19/England/ALDP-97CCE4/2020\|EPI_ISL_526316\|20 | ···················· | ···················· | ····················· | ···················· | ···················· | ···················· |
| G | 716 hCoV-19/England/ALDP-97CD0F/2020\|EPI_ISL_526317\|20 | ···················· | ···················· | ····················· | ···················· | ···················· | ···················· |
| G | 717 hCoV-19/England/ALDP-97CF90/2020\|EPI_ISL_526318\|20 | ···················· | ···················· | ····················· | ···················· | ···················· | ···················· |
| G | 718 hCoV-19/England/ALDP-97CFAF/2020\|EPI_ISL_526319\|20 | ···················· | ···················· | ····················· | ···················· | ···················· | ···················· |
| G | 719 hCoV-19/England/ALDP-97D357/2020\|EPI_ISL_526323\|20 | ···················· | ···················· | ····················· | ···················· | ···················· | ···················· |
| G | 720 hCoV-19/England/ALDP-97D384/2020\|EPI_ISL_526324\|20 | ···················· | ···················· | ····················· | ···················· | ···················· | ···················· |
| G | 721 hCoV-19/England/ALDP-97D393/2020\|EPI_ISL_526325\|20 | ···················· | ···················· | ····················· | ···················· | ···················· | ···················· |
| G | 722 hCoV-19/England/ALDP-97D6A9/2020\|EPI_ISL_526327\|20 | ···················· | ···················· | ····················· | ···················· | ···················· | ···················· |
| G | 723 hCoV-19/England/ALDP-97DDB3/2020\|EPI_ISL_526333\|20 | ···················· | ···················· | ····················· | ···················· | ···················· | ···················· |
| G | 724 hCoV-19/England/ALDP-97DE0B/2020\|EPI_ISL_526334\|20 | ···················· | ···················· | ····················· | ···················· | ···················· | ···················· |
| G | 725 hCoV-19/England/ALDP-97E040/2020\|EPI_ISL_526336\|20 | ···················· | ···················· | ····················· | ···················· | ···················· | ···················· |
| G | 726 hCoV-19/England/ALDP-97F7C2/2020\|EPI_ISL_526337\|20 | ···················· | ·A·················· | ····················· | ···················· | ···················· | ··A················· |
| G | 727 hCoV-19/England/ALDP-97F80B/2020\|EPI_ISL_526338\|20 | ···················· | ···················· | ····················· | ···················· | ···················· | ···················· |
| G | 728 hCoV-19/England/ALDP-97FA32/2020\|EPI_ISL_526339\|20 | ···················· | ···················· | ····················· | ···················· | ···················· | ···················· |
| G | 729 hCoV-19/England/LIVE-A975F/2020\|EPI_ISL_526390\|202 | ···················· | ···················· | ····················· | ···················· | ···················· | ···················· |
| G | 730 hCoV-19/England/LIVE-A976E/2020\|EPI_ISL_526391\|202 | ···················· | ···················· | ····················· | ···················· | ···················· | ···················· |
| G | 731 hCoV-19/England/LIVE-A979B/2020\|EPI_ISL_526393\|202 | ···················· | ···················· | ····················· | ···················· | ···················· | ···················· |
| G | 732 hCoV-19/England/LIVE-A97AA/2020\|EPI_ISL_526394\|202 | ···················· | ···················· | ····················· | ···················· | ···················· | ···················· |
| G | 733 hCoV-19/England/MILK-9772C9/2020\|EPI_ISL_526395\|20 | ···················· | ···················· | ····················· | ···················· | ···················· | ···················· |
| G | 734 hCoV-19/England/MILK-9773C6/2020\|EPI_ISL_526396\|20 | ···················· | ···················· | ····················· | ···················· | ···················· | ···················· |
| G | 735 hCoV-19/England/MILK-97742D/2020\|EPI_ISL_526397\|20 | ···················· | ···················· | ····················· | ···················· | ···················· | ···················· |
| G | 736 hCoV-19/England/MILK-97743C/2020\|EPI_ISL_526398\|20 | ···················· | ···················· | ····················· | ···················· | ···················· | ···················· |
| G | 737 hCoV-19/England/MILK-977663/2020\|EPI_ISL_526399\|20 | ···················· | ···················· | ····················· | ···················· | ···················· | ···················· |
| G | 738 hCoV-19/USA/AR-CDC-4221/2020\|EPI_ISL_527639\|2020-0 | ···················· | ···················· | ····················· | ···················· | ···················· | ···················· |
| G | 739 hCoV-19/USA/AR-CDC-4226/2020\|EPI_ISL_527640\|2020-0 | ···················· | ···················· | ····················· | ···················· | ···················· | ···················· |
| G | 740 hCoV-19/USA/AR-CDC-4227/2020\|EPI_ISL_527641\|2020-0 | ···················· | ···················· | ····················· | ···················· | ···················· | ···················· |
| G | 741 hCoV-19/USA/AR-CDC-4253/2020\|EPI_ISL_527642\|2020-0 | ···················· | ···················· | ····················· | ···················· | ···················· | ···················· |
| G | 742 hCoV-19/USA/AR-CDC-4265/2020\|EPI_ISL_527643\|2020-0 | ···················· | ···················· | ····················· | ···················· | ···················· | ···················· |
| G | 743 hCoV-19/USA/AR-CDC-4270/2020\|EPI_ISL_527644\|2020-0 | ···················· | ···················· | ····················· | ···················· | ···················· | ···················· |
| G | 744 hCoV-19/USA/AR-CDC-4271/2020\|EPI_ISL_527645\|2020-0 | ···················· | ···················· | ····················· | ···················· | ···················· | ···················· |
| G | 745 hCoV-19/USA/AR-CDC-4288/2020\|EPI_ISL_527646\|2020-0 | ···················· | ···················· | ····················· | ···················· | ···················· | ···················· |
| G | 746 hCoV-19/USA/AR-CDC-4296/2020\|EPI_ISL_527647\|2020-0 | ···················· | ···················· | ····················· | ···················· | ···················· | ···················· |
| G | 747 hCoV-19/USA/AR-CDC-4299/2020\|EPI_ISL_527648\|2020-0 | ···················· | ···················· | ····················· | ···················· | ···················· | ···················· |
| G | 748 hCoV-19/USA/AR-CDC-4307/2020\|EPI_ISL_527649\|2020-0 | ···················· | ···················· | ····················· | ···················· | ···················· | ···················· |
| G | 749 hCoV-19/USA/AR-CDC-4321/2020\|EPI_ISL_527650\|2020-0 | ···················· | ···················· | ····················· | ···················· | ···················· | ···················· |
| G | 750 hCoV-19/USA/AR-CDC-4373/2020\|EPI_ISL_527651\|2020-0 | ···················· | ···················· | ····················· | ···················· | ···················· | ···················· |
| G | 751 hCoV-19/USA/AR-CDC-4374/2020\|EPI_ISL_527652\|2020-0 | ···················· | ···················· | ····················· | ···················· | ···················· | ···················· |
| G | 752 hCoV-19/USA/AR-CDC-4375/2020\|EPI_ISL_527653\|2020-0 | ···················· | ···················· | ····················· | ···················· | ···················· | ···················· |
| G | 753 hCoV-19/USA/AR-CDC-4378/2020\|EPI_ISL_527654\|2020-0 | ···················· | ···················· | ····················· | ···················· | ···················· | ···················· |
| G | 754 hCoV-19/USA/AR-CDC-4380/2020\|EPI_ISL_527655\|2020-0 | ···················· | ···················· | ····················· | ···················· | ···················· | ···················· |
| G | 755 hCoV-19/USA/AR-CDC-4388/2020\|EPI_ISL_527656\|2020-0 | ···················· | ···················· | ····················· | ···················· | ···················· | ···················· |
| G | 756 hCoV-19/USA/AR-CDC-6567/2020\|EPI_ISL_527657\|2020-0 | ···················· | ···················· | ····················· | ···················· | ···················· | ···················· |
| G | 757 hCoV-19/USA/GA-CDC-6258/2020\|EPI_ISL_527658\|2020-0 | ···················· | ···················· | ····················· | ···················· | ···················· | ···················· |
| G | 758 hCoV-19/USA/GA-CDC-6259/2020\|EPI_ISL_527659\|2020-0 | ···················· | ···················· | ····················· | ···················· | ···················· | ···················· |
| G | 759 hCoV-19/USA/GA-CDC-6435/2020\|EPI_ISL_527664\|2020-0 | ···················· | ···················· | ····················· | ···················· | ···················· | ···················· |
| G | 760 hCoV-19/USA/GA-CDC-6438/2020\|EPI_ISL_527665\|2020-0 | ···················· | ···················· | ····················· | ···················· | ···················· | ···················· |
| G | 761 hCoV-19/USA/IN-CDC-6387/2020\|EPI_ISL_527666\|2020-0 | ···················· | ···················· | ····················· | ···················· | ···················· | ···················· |
| G | 762 hCoV-19/USA/MN-CDC-6186/2020\|EPI_ISL_527667\|2020-0 | ···················· | ···················· | ····················· | ···················· | ···················· | ···················· |
| G | 763 hCoV-19/USA/MN-CDC-6189/2020\|EPI_ISL_527668\|2020-0 | ···················· | ···················· | ····················· | ···················· | ···················· | ···················· |

| | | | | | | | | |
|---|---|---|---|---|---|---|---|---|
| G | 764 | hCoV-19/Italy/ABR-IZSGC-TE46424/2020\|EPI_ISL_52899 | ···················· | ···················· | ···················· | ···················· | ···················· | ···················· |
| G | 765 | hCoV-19/Italy/ABR-IZSGC-TE46428/2020\|EPI_ISL_52899 | ···················· | ···················· | ···················· | ···················· | ···················· | ···················· |
| G | 766 | hCoV-19/Italy/ABR-IZSGC-TE46429/2020\|EPI_ISL_52899 | ···················· | ···················· | ···················· | ···················· | ···················· | ···················· |
| G | 767 | hCoV-19/Italy/ABR-IZSGC-TE46433/2020\|EPI_ISL_52899 | ···················· | ···················· | ···················· | ···················· | ···················· | ···················· |
| G | 768 | hCoV-19/Italy/ABR-IZSGC-TE46435/2020\|EPI_ISL_52899 | ···················· | ···················· | ···················· | ···················· | ···················· | ···················· |
| G | 769 | hCoV-19/Singapore/826/2020\|EPI_ISL_516826\|2020-08- | ···················· | ···················· | ···················· | ···················· | ···················· | ···················· |
| G | 770 | hCoV-19/Indonesia/YO-UGM-781481/2020\|EPI_ISL_51682 | ················G··· | ···················· | ···················· | ···················· | ···················· | ···················· |
| G | 771 | hCoV-19/England/LOND-1260133/2020\|EPI_ISL_516841\|2 | ···················· | ···················· | ···················· | ···················· | ···················· | ···················· |
| G | 772 | hCoV-19/England/LOND-125FCAF/2020\|EPI_ISL_516842\|2 | ···················· | ···················· | ···················· | ···················· | ···················· | ···················· |
| G | 773 | hCoV-19/England/LOND-1260A47/2020\|EPI_ISL_516843\|2 | ···················· | ···················· | ···················· | ···················· | ···················· | ···················· |
| G | 774 | hCoV-19/England/LOND-12605FB/2020\|EPI_ISL_516844\|2 | ···················· | ···················· | ···················· | ···················· | ···················· | ···················· |
| G | 775 | hCoV-19/England/LOND-1260A38/2020\|EPI_ISL_516846\|2 | ···T················ | ···················· | ···················· | ···················· | ···················· | ···················· |
| G | 776 | hCoV-19/England/LOND-125FEF4/2020\|EPI_ISL_516848\|2 | ···················· | ···················· | ···················· | ···················· | ···················· | ···················· |
| G | 777 | hCoV-19/England/LOND-1260124/2020\|EPI_ISL_516852\|2 | ···················· | ···················· | ···················· | ···················· | ···················· | ···················· |
| G | 778 | hCoV-19/England/LOND-12606DA/2020\|EPI_ISL_516857\|2 | ···················· | ···················· | ···················· | ···················· | ···················· | ···················· |
| G | 779 | hCoV-19/England/LOND-12609D1/2020\|EPI_ISL_516858\|2 | ···················· | ···················· | ···················· | ···················· | ···················· | ···················· |
| G | 780 | hCoV-19/England/LOND-126090E/2020\|EPI_ISL_516863\|2 | ···················· | ···················· | ···················· | ···················· | ···················· | ···················· |
| G | 781 | hCoV-19/England/LOND-125FFC4/2020\|EPI_ISL_516864\|2 | ···················· | ···················· | ···················· | ···················· | ···················· | ···················· |
| G | 782 | hCoV-19/England/LOND-125FF79/2020\|EPI_ISL_516865\|2 | ···················· | ···················· | ···················· | ···················· | ···················· | ···················· |
| G | 783 | hCoV-19/England/LOND-12609A4/2020\|EPI_ISL_516866\|2 | ···················· | ···················· | ···················· | ···················· | ···················· | ···················· |
| G | 784 | hCoV-19/England/LOND-1260018/2020\|EPI_ISL_516868\|2 | ···················· | ···················· | ···················· | ···················· | ···················· | ···················· |
| G | 785 | hCoV-19/Australia/VIC4599/2020\|EPI_ISL_519000\|2020 | ···················· | ···················· | ···················· | ···················· | ···················· | ···················· |
| G | 786 | hCoV-19/Australia/VIC4600/2020\|EPI_ISL_519001\|2020 | ···················· | ···················· | ···················· | ···················· | ···················· | ···················· |
| G | 787 | hCoV-19/Australia/VIC4601/2020\|EPI_ISL_519002\|2020 | ···················· | ···················· | ···················· | ···················· | ···················· | ···················· |
| G | 788 | hCoV-19/Australia/VIC4602/2020\|EPI_ISL_519003\|2020 | ···················· | ···················· | ···················· | ···················· | ···················· | ···················· |
| G | 789 | hCoV-19/Australia/VIC4603/2020\|EPI_ISL_519004\|2020 | ···················· | ···················· | ···················· | ···················· | ···················· | ···················· |
| G | 790 | hCoV-19/Australia/VIC4605/2020\|EPI_ISL_519006\|2020 | ···················· | ···················· | ···················· | ···················· | ···················· | ···················· |
| G | 791 | hCoV-19/Australia/VIC4607/2020\|EPI_ISL_519008\|2020 | ···················· | ···················· | ···················· | ···················· | ···················· | ···················· |
| G | 792 | hCoV-19/Australia/VIC4608/2020\|EPI_ISL_519009\|2020 | ···················· | ···················· | ···················· | ···················· | ···················· | ···················· |
| G | 793 | hCoV-19/Australia/VIC4609/2020\|EPI_ISL_519010\|2020 | ···················· | ···················· | ···················· | ···················· | ···················· | ···················· |
| G | 794 | hCoV-19/Australia/VIC4610/2020\|EPI_ISL_519011\|2020 | ···················· | ···················· | ···················· | ···················· | ···················· | ···················· |
| G | 795 | hCoV-19/Australia/VIC4615/2020\|EPI_ISL_519013\|2020 | ···················· | ···················· | ···················· | ···················· | ···················· | ···················· |
| G | 796 | hCoV-19/Australia/VIC4617/2020\|EPI_ISL_519014\|2020 | ···················· | ···················· | ···················· | ···················· | ···················· | ···················· |
| G | 797 | hCoV-19/Australia/VIC4619/2020\|EPI_ISL_519015\|2020 | ···················· | ···················· | ···················· | ···················· | ···················· | ···················· |
| G | 798 | hCoV-19/Australia/VIC4620/2020\|EPI_ISL_519016\|2020 | ···················· | ···················· | ···················· | ···················· | ···················· | ···················· |
| G | 799 | hCoV-19/Australia/VIC4621/2020\|EPI_ISL_519017\|2020 | ···················· | ···················· | ···················· | ···················· | ···················· | ···················· |
| G | 800 | hCoV-19/Australia/VIC4622/2020\|EPI_ISL_519018\|2020 | ···················· | ···················· | ···················· | ···················· | ···················· | ···················· |
| G | 801 | hCoV-19/Australia/VIC4625/2020\|EPI_ISL_519019\|2020 | ···················· | ···················· | ···················· | ···················· | ···················· | ···················· |
| G | 802 | hCoV-19/Australia/VIC4627/2020\|EPI_ISL_519020\|2020 | ···················· | ···················· | ···················· | ···················· | ···················· | ···················· |
| G | 803 | hCoV-19/Australia/VIC4629/2020\|EPI_ISL_519022\|2020 | ···················· | ···················· | ···················· | ···················· | ···················· | ···················· |
| G | 804 | hCoV-19/Australia/VIC4632/2020\|EPI_ISL_519023\|2020 | ···················· | ···················· | ···················· | ···················· | ···················· | ···················· |
| G | 805 | hCoV-19/Australia/VIC4634/2020\|EPI_ISL_519025\|2020 | ···················· | ···················· | ···················· | ···················· | ···················· | ···················· |
| G | 806 | hCoV-19/Australia/VIC4635/2020\|EPI_ISL_519026\|2020 | ···················· | ···················· | ···················· | ···················· | ···················· | ···················· |
| G | 807 | hCoV-19/Australia/VIC4636/2020\|EPI_ISL_519027\|2020 | ···················· | ···················· | ···················· | ···················· | ···················· | ···················· |
| G | 808 | hCoV-19/Australia/VIC4637/2020\|EPI_ISL_519028\|2020 | ···················· | ···················· | ···················· | ···················· | ···················· | ···················· |
| G | 809 | hCoV-19/Australia/VIC4639/2020\|EPI_ISL_519030\|2020 | ···················· | ···················· | ···················· | ···················· | ···················· | ···················· |
| G | 810 | hCoV-19/Australia/VIC4640/2020\|EPI_ISL_519031\|2020 | ···················· | ···················· | ···················· | ···················· | ···················· | ···················· |
| G | 811 | hCoV-19/Australia/VIC4641/2020\|EPI_ISL_519032\|2020 | ···················· | ···················· | ···················· | ···················· | ···················· | ···················· |
| G | 812 | hCoV-19/Australia/VIC4642/2020\|EPI_ISL_519033\|2020 | ···················· | ···················· | ···················· | ···················· | ···················· | ···················· |
| G | 813 | hCoV-19/Australia/VIC4643/2020\|EPI_ISL_519034\|2020 | ···················· | ···················· | ···················· | ···················· | ···················· | ···················· |
| G | 814 | hCoV-19/Australia/VIC4644/2020\|EPI_ISL_519035\|2020 | ···················· | ···················· | ···················· | ···················· | ···················· | ···················· |
| G | 815 | hCoV-19/Australia/VIC4645/2020\|EPI_ISL_519036\|2020 | ···················· | ···················· | ···················· | ···················· | ···················· | ···················· |
| G | 816 | hCoV-19/Australia/VIC4646/2020\|EPI_ISL_519037\|2020 | ···················· | ···················· | ···················· | ···················· | ···················· | ···················· |
| G | 817 | hCoV-19/Australia/VIC4647/2020\|EPI_ISL_519038\|2020 | ···················· | ···················· | ···················· | ···················· | ···················· | ···················· |
| G | 818 | hCoV-19/Australia/VIC4648/2020\|EPI_ISL_519039\|2020 | ···················· | ···················· | ···················· | ···················· | ···················· | ···················· |
| G | 819 | hCoV-19/Australia/VIC4654/2020\|EPI_ISL_519043\|2020 | ···················· | ···················· | ···················· | ···················· | ···················· | ···················· |
| G | 820 | hCoV-19/Australia/VIC4655/2020\|EPI_ISL_519044\|2020 | ···················· | ···················· | ···················· | ···················· | ···················· | ···················· |
| G | 821 | hCoV-19/Australia/VIC4656/2020\|EPI_ISL_519045\|2020 | ···················· | ···················· | ···················· | ···················· | ···················· | ···················· |
| G | 822 | hCoV-19/Australia/VIC4657/2020\|EPI_ISL_519046\|2020 | ···················· | ···················· | ···················· | ···················· | ···················· | ···················· |
| G | 823 | hCoV-19/Australia/VIC4658/2020\|EPI_ISL_519047\|2020 | ···················· | ···················· | ···················· | ···················· | ···················· | ···················· |
| G | 824 | hCoV-19/Australia/VIC4659/2020\|EPI_ISL_519048\|2020 | ···················· | ···················· | ···················· | ···················· | ···················· | ···················· |
| G | 825 | hCoV-19/Australia/VIC4660/2020\|EPI_ISL_519049\|2020 | ···················· | ···················· | ···················· | ···················· | ···················· | ···················· |
| G | 826 | hCoV-19/Australia/VIC5235/2020\|EPI_ISL_520000\|2020 | ···················· | ···················· | ···················· | ···················· | ···················· | ···················· |
| G | 827 | hCoV-19/Australia/VIC5236/2020\|EPI_ISL_520001\|2020 | ···················· | ···················· | ···················· | ···················· | ···················· | ···················· |
| G | 828 | hCoV-19/Australia/VIC5237/2020\|EPI_ISL_520002\|2020 | ···················· | ···················· | ···················· | ···················· | ···················· | ···················· |
| G | 829 | hCoV-19/Australia/VIC5238/2020\|EPI_ISL_520003\|2020 | ···················· | ···················· | ···················· | ···················· | ···················· | ···················· |
| G | 830 | hCoV-19/Australia/VIC5239/2020\|EPI_ISL_520004\|2020 | ···················· | ···················· | ···················· | ···················· | ···················· | ···················· |
| G | 831 | hCoV-19/Australia/VIC5240/2020\|EPI_ISL_520005\|2020 | ···················· | ···················· | ···················· | ···················· | ···················· | ···················· |
| G | 832 | hCoV-19/Australia/VIC5241/2020\|EPI_ISL_520006\|2020 | ···················· | ···················· | ···················· | ···················· | ···················· | ···················· |
| G | 833 | hCoV-19/Australia/VIC5242/2020\|EPI_ISL_520007\|2020 | ···················· | ···················· | ···················· | ···················· | ···················· | ···················· |
| G | 834 | hCoV-19/Australia/VIC5243/2020\|EPI_ISL_520008\|2020 | ···················· | ···················· | ···················· | ···················· | ···················· | ···················· |
| G | 835 | hCoV-19/Australia/VIC5244/2020\|EPI_ISL_520009\|2020 | ···················· | ···················· | ···················· | ···················· | ···················· | ···················· |
| G | 836 | hCoV-19/Australia/VIC5245/2020\|EPI_ISL_520010\|2020 | ···················· | ···················· | ···················· | ···················· | ···················· | ···················· |
| G | 837 | hCoV-19/Australia/VIC5246/2020\|EPI_ISL_520011\|2020 | ···················· | ···················· | ···················· | ···················· | ···················· | ···················· |
| G | 838 | hCoV-19/Australia/VIC5247/2020\|EPI_ISL_520012\|2020 | ···················· | ···················· | ···················· | ···················· | ···················· | ···················· |
| G | 839 | hCoV-19/Australia/VIC5248/2020\|EPI_ISL_520013\|2020 | ···················· | ···················· | ···················· | ···················· | ···················· | ···················· |
| G | 840 | hCoV-19/Australia/VIC5249/2020\|EPI_ISL_520014\|2020 | ···················· | ···················· | ···················· | ···················· | ···················· | ···················· |
| G | 841 | hCoV-19/Australia/VIC5250/2020\|EPI_ISL_520015\|2020 | ···················· | ···················· | ···················· | ···················· | ···················· | ···················· |
| G | 842 | hCoV-19/Australia/VIC5251/2020\|EPI_ISL_520016\|2020 | ···················· | ···················· | ···················· | ···················· | ···················· | ···················· |
| G | 843 | hCoV-19/Australia/VIC5252/2020\|EPI_ISL_520017\|2020 | ···················· | ···················· | ···················· | ···················· | ···················· | ···················· |

| | | | | | | | | |
|---|---|---|---|---|---|---|---|---|
| G | 844 hCoV-19/Australia/VIC5253/2020\|EPI_ISL_520018\|2020 | ···················· | ···················· | ···················· | ···················· | ···················· | ···················· | |
| G | 845 hCoV-19/Australia/VIC5254/2020\|EPI_ISL_520019\|2020 | ···················· | ···················· | ···················· | ···················· | ···················· | ···················· | |
| G | 846 hCoV-19/Australia/VIC5255/2020\|EPI_ISL_520020\|2020 | ···················· | ···················· | ···················· | ···················· | ···················· | ···················· | |
| G | 847 hCoV-19/Australia/VIC5256/2020\|EPI_ISL_520021\|2020 | ···················· | ···················· | ···················· | ···················· | ···················· | ···················· | |
| G | 848 hCoV-19/Australia/VIC5257/2020\|EPI_ISL_520022\|2020 | ···················· | ···················· | ···················· | ···················· | ···················· | ···················· | |
| G | 849 hCoV-19/Australia/VIC5258/2020\|EPI_ISL_520023\|2020 | ···················· | ···················· | ···················· | ···················· | ···················· | ···················· | |
| G | 850 hCoV-19/Australia/VIC5259/2020\|EPI_ISL_520024\|2020 | ···················· | ···················· | ···················· | ···················· | ···················· | ···················· | |
| G | 851 hCoV-19/Australia/VIC5260/2020\|EPI_ISL_520025\|2020 | ···················· | ···················· | ···················· | ···················· | ···················· | ···················· | |
| G | 852 hCoV-19/Australia/VIC5261/2020\|EPI_ISL_520026\|2020 | ···················· | ···················· | ···················· | ···················· | ···················· | ···················· | |
| G | 853 hCoV-19/Australia/VIC5327/2020\|EPI_ISL_520090\|2020 | ···················· | ···················· | ···················· | ···················· | ···················· | ···················· | |
| G | 854 hCoV-19/Australia/VIC5328/2020\|EPI_ISL_520091\|2020 | ···················· | ···················· | ···················· | ···················· | ···················· | ···················· | |
| G | 855 hCoV-19/Australia/VIC5329/2020\|EPI_ISL_520092\|2020 | ···················· | ···················· | ···················· | ···················· | ···················· | ···················· | |
| G | 856 hCoV-19/Australia/VIC5330/2020\|EPI_ISL_520093\|2020 | ···················· | ···················· | ···················· | ···················· | ···················· | ···················· | |
| G | 857 hCoV-19/Australia/VIC5331/2020\|EPI_ISL_520094\|2020 | ···················· | ···················· | ···················· | ···················· | ···················· | ···················· | |
| G | 858 hCoV-19/Australia/VIC5332/2020\|EPI_ISL_520095\|2020 | ···················· | ···················· | ···················· | ···················· | ···················· | ···················· | |
| G | 859 hCoV-19/Australia/VIC5333/2020\|EPI_ISL_520096\|2020 | ···················· | ···················· | ···················· | ···················· | ···················· | ···················· | |
| G | 860 hCoV-19/Australia/VIC5334/2020\|EPI_ISL_520097\|2020 | ···················· | ···················· | ···················· | ···················· | ···················· | ···················· | |
| G | 861 hCoV-19/Australia/VIC5335/2020\|EPI_ISL_520098\|2020 | ···················· | ···················· | ···················· | ···················· | ···················· | ···················· | |
| G | 862 hCoV-19/Australia/VIC5336/2020\|EPI_ISL_520099\|2020 | ···················· | ···················· | ···················· | ···················· | ···················· | ···················· | |
| G | 863 hCoV-19/Australia/VIC6530/2020\|EPI_ISL_521300\|2020 | ···················· | ···················· | ···················· | ···················· | ···················· | ···················· | |
| G | 864 hCoV-19/Australia/VIC6531/2020\|EPI_ISL_521301\|2020 | ···················· | ···················· | ···················· | ···················· | ···················· | ···················· | |
| G | 865 hCoV-19/Australia/VIC6532/2020\|EPI_ISL_521302\|2020 | ···················· | ···················· | ···················· | ···················· | ···················· | ···················· | |
| G | 866 hCoV-19/Australia/VIC6533/2020\|EPI_ISL_521303\|2020 | ···················· | ···················· | ···················· | ···················· | ···················· | ···················· | |
| G | 867 hCoV-19/Australia/VIC6534/2020\|EPI_ISL_521304\|2020 | ···················· | ···················· | ···················· | ···················· | ···················· | ···················· | |
| G | 868 hCoV-19/Australia/VIC6535/2020\|EPI_ISL_521305\|2020 | ···················· | ···················· | ···················· | ···················· | ···················· | ···················· | |
| G | 869 hCoV-19/Australia/VIC6536/2020\|EPI_ISL_521306\|2020 | ···················· | ···················· | ···················· | ···················· | ···················· | ···················· | |
| G | 870 hCoV-19/Australia/VIC6537/2020\|EPI_ISL_521307\|2020 | ···················· | ···················· | ···················· | ···················· | ···················· | ···················· | |
| G | 871 hCoV-19/Australia/VIC6538/2020\|EPI_ISL_521308\|2020 | ···················· | ···················· | ···················· | ···················· | ···················· | ···················· | |
| G | 872 hCoV-19/Australia/VIC6539/2020\|EPI_ISL_521309\|2020 | ···················· | ···················· | ···················· | ···················· | ···················· | ···················· | |
| G | 873 hCoV-19/Australia/VIC6540/2020\|EPI_ISL_521310\|2020 | ···················· | ···················· | ···················· | ···················· | ···················· | ···················· | |
| G | 874 hCoV-19/Australia/VIC6541/2020\|EPI_ISL_521311\|2020 | ···················· | ···················· | ···················· | ···················· | ···················· | ···················· | |
| G | 875 hCoV-19/Australia/VIC6542/2020\|EPI_ISL_521312\|2020 | ···················· | ···················· | ···················· | ···················· | ···················· | ···················· | |
| G | 876 hCoV-19/Australia/VIC6543/2020\|EPI_ISL_521313\|2020 | ···················· | ···················· | ···················· | ···················· | ···················· | ···················· | |
| G | 877 hCoV-19/Australia/VIC6544/2020\|EPI_ISL_521314\|2020 | ···················· | ···················· | ···················· | ···················· | ···················· | ···················· | |
| G | 878 hCoV-19/Australia/VIC6545/2020\|EPI_ISL_521315\|2020 | ···················· | ···················· | ···················· | ···················· | ···················· | ···················· | |
| G | 879 hCoV-19/Australia/VIC6546/2020\|EPI_ISL_521316\|2020 | ···················· | ···················· | ···················· | ···················· | ···················· | ···················· | |
| G | 880 hCoV-19/Australia/VIC6547/2020\|EPI_ISL_521317\|2020 | ···················· | ···················· | ···················· | ···················· | ···················· | ···················· | |
| G | 881 hCoV-19/Australia/VIC6549/2020\|EPI_ISL_521318\|2020 | ···················· | ···················· | ···················· | ···················· | ···················· | ···················· | |
| G | 882 hCoV-19/Australia/VIC6552/2020\|EPI_ISL_521320\|2020 | ···················· | ···················· | ···················· | ···················· | ···················· | ···················· | |
| G | 883 hCoV-19/Australia/VIC6553/2020\|EPI_ISL_521321\|2020 | ···················· | ···················· | ···················· | ···················· | ···················· | ···················· | |
| G | 884 hCoV-19/Australia/VIC6557/2020\|EPI_ISL_521325\|2020 | ···················· | ···················· | ···················· | ···················· | ···················· | ···················· | |
| G | 885 hCoV-19/Australia/VIC6558/2020\|EPI_ISL_521326\|2020 | ···················· | ···················· | ···················· | ···················· | ···················· | ···················· | |
| G | 886 hCoV-19/Australia/VIC6560/2020\|EPI_ISL_521327\|2020 | ···················· | ···················· | ···················· | ···················· | ···················· | ···················· | |
| G | 887 hCoV-19/Australia/VIC6561/2020\|EPI_ISL_521328\|2020 | ···················· | ···················· | ···················· | ···················· | ···················· | ···················· | |
| G | 888 hCoV-19/Australia/VIC6563/2020\|EPI_ISL_521329\|2020 | ···················· | ···················· | ···················· | ···················· | ···················· | ···················· | |
| G | 889 hCoV-19/Australia/VIC6564/2020\|EPI_ISL_521330\|2020 | ···················· | ···················· | ···················· | ···················· | ···················· | ···················· | |
| G | 890 hCoV-19/Australia/VIC6565/2020\|EPI_ISL_521331\|2020 | ···················· | ···················· | ···················· | ···················· | ···················· | ···················· | |
| G | 891 hCoV-19/Australia/VIC6566/2020\|EPI_ISL_521332\|2020 | ···················· | ···················· | ···················· | ···················· | ···················· | ···················· | |
| G | 892 hCoV-19/Australia/VIC6568/2020\|EPI_ISL_521334\|2020 | ···················· | ···················· | ···················· | ···················· | ···················· | ···················· | |
| G | 893 hCoV-19/Australia/VIC6569/2020\|EPI_ISL_521335\|2020 | ···················· | ···················· | ···················· | ···················· | ···················· | ···················· | |
| G | 894 hCoV-19/Australia/VIC6572/2020\|EPI_ISL_521338\|2020 | ···················· | ···················· | ···················· | ···················· | ···················· | ···················· | |
| G | 895 hCoV-19/Australia/VIC6573/2020\|EPI_ISL_521339\|2020 | ···················· | ···················· | ···················· | ···················· | ···················· | ···················· | |
| G | 896 hCoV-19/Australia/VIC6575/2020\|EPI_ISL_521340\|2020 | ···················· | ···················· | ···················· | ···················· | ···················· | ···················· | |
| G | 897 hCoV-19/Australia/VIC6577/2020\|EPI_ISL_521341\|2020 | ···················· | ···················· | ···················· | ···················· | ···················· | ···················· | |
| G | 898 hCoV-19/Australia/VIC6578/2020\|EPI_ISL_521342\|2020 | ···················· | ···················· | ···················· | ···················· | ···················· | ···················· | |
| G | 899 hCoV-19/Australia/VIC6579/2020\|EPI_ISL_521343\|2020 | ···················· | ···················· | ···················· | ···················· | ···················· | ···················· | |
| G | 900 hCoV-19/Australia/VIC6581/2020\|EPI_ISL_521345\|2020 | ···················· | ···················· | ···················· | ···················· | ···················· | ···················· | |
| G | 901 hCoV-19/Australia/VIC6582/2020\|EPI_ISL_521346\|2020 | ···················· | ···················· | ···················· | ···················· | ···················· | ···················· | |
| G | 902 hCoV-19/Australia/VIC6583/2020\|EPI_ISL_521347\|2020 | ···················· | ···················· | ···················· | ···················· | ···················· | ···················· | |
| G | 903 hCoV-19/Australia/VIC6585/2020\|EPI_ISL_521348\|2020 | ···················· | ···················· | ···················· | ···················· | ···················· | ···················· | |
| G | 904 hCoV-19/Australia/VIC6586/2020\|EPI_ISL_521349\|2020 | ···················· | ···················· | ···················· | ···················· | ···················· | ···················· | |
| G | 905 hCoV-19/Australia/VIC6587/2020\|EPI_ISL_521350\|2020 | ···················· | ···················· | ···················· | ···················· | ···················· | ···················· | |
| G | 906 hCoV-19/Australia/VIC6590/2020\|EPI_ISL_521352\|2020 | ···················· | ···················· | ···················· | ···················· | ···················· | ···················· | |
| G | 907 hCoV-19/Australia/VIC6591/2020\|EPI_ISL_521353\|2020 | ···················· | ···················· | ···················· | ···················· | ···················· | ···················· | |
| G | 908 hCoV-19/Australia/VIC6592/2020\|EPI_ISL_521354\|2020 | ···················· | ···················· | ···················· | ···················· | ···················· | ···················· | |
| G | 909 hCoV-19/Australia/VIC6593/2020\|EPI_ISL_521355\|2020 | ···················· | ···················· | ···················· | ···················· | ···················· | ···················· | |
| G | 910 hCoV-19/Australia/VIC6594/2020\|EPI_ISL_521356\|2020 | ···················· | ···················· | ···················· | ···················· | ···················· | ···················· | |
| G | 911 hCoV-19/Australia/VIC6595/2020\|EPI_ISL_521357\|2020 | ···················· | ···················· | ···················· | ···················· | ···················· | ···················· | |
| G | 924 hCoV-19/Australia/TAS236/2020\|EPI_ISL_522665\|2020- | ···················· | ···················· | ···················· | ···················· | ···················· | ···················· | |
| G | 928 hCoV-19/Australia/TAS243/2020\|EPI_ISL_522671\|2020- | ···················· | ···················· | ···················· | ···················· | ···················· | ···················· | |
| G | 930 hCoV-19/Australia/TAS246/2020\|EPI_ISL_522673\|2020- | ···················· | ···················· | ···················· | ···················· | ···················· | ···················· | |
| G | 934 hCoV-19/Australia/TAS250/2020\|EPI_ISL_522678\|2020- | ···················· | ···················· | ···················· | ···················· | NNN················· | ···················· | 1 |
| G | 935 hCoV-19/Australia/TAS251/2020\|EPI_ISL_522679\|2020- | ···················· | ···················· | ···················· | ···················· | ···················· | ···················· | |
| G | 936 hCoV-19/Australia/TAS252/2020\|EPI_ISL_522680\|2020- | ···················· | ···················· | ···················· | ···················· | ···················· | ···················· | |
| G | 938 hCoV-19/Australia/TAS255/2020\|EPI_ISL_522683\|2020- | ···················· | ···················· | ···················· | ···················· | ···················· | ···················· | |
| G | 939 hCoV-19/Australia/TAS258/2020\|EPI_ISL_522685\|2020 | ···················· | ···················· | ···················· | ···················· | ···················· | ···················· | |
| G | 940 hCoV-19/Peru/LAM-INS-071/2020\|EPI_ISL_523954\|2020- | ···················· | ···················· | ···················· | ···················· | ···················· | ···················· | |
| G | 941 hCoV-19/Brazil/SP-128/2020\|EPI_ISL_523955\|2020-04- | ···················· | ···················· | ···················· | ···················· | ···················· | ···················· | |
| G | 942 hCoV-19/Brazil/SP-132/2020\|EPI_ISL_523956\|2020-03- | ···················· | ···················· | ···················· | ···················· | ···················· | ···················· | |
| G | 943 hCoV-19/Brazil/SP-133/2020\|EPI_ISL_523957\|2020-04- | ···················· | ···················· | ···················· | ···················· | ···················· | ···················· | |

| | | | | | | | | |
|---|---|---|---|---|---|---|---|---|
| G | 944 | hCoV-19/Brazil/SP-162/2020\|EPI_ISL_523958\|2020-04- | .................... | .................... | .................... | .................... | .................... | .................... |
| G | 945 | hCoV-19/Brazil/SP-163/2020\|EPI_ISL_523959\|2020-08- | .................... | .................... | .................... | .................... | .................... | .................... |
| G | 946 | hCoV-19/Moldova/ChVir7768/2020\|EPI_ISL_523960\|2020 | .................... | .................... | .................... | .................... | .................... | .................... |
| G | 947 | hCoV-19/Brazil/SP-164/2020\|EPI_ISL_523961\|2020-04- | .................... | .................... | .................... | .................... | .................... | .................... |
| G | 948 | hCoV-19/Moldova/ChVir7770/2020\|EPI_ISL_523962\|2020 | .................... | .................... | .................... | .................... | .................... | .................... |
| G | 949 | hCoV-19/Brazil/SP-165/2020\|EPI_ISL_523963\|2020-03- | .................... | .................... | .................... | .................... | .................... | .................... |
| G | 950 | hCoV-19/Moldova/ChVir7771/2020\|EPI_ISL_523964\|2020 | .................... | .................... | .................... | .................... | .................... | .................... |
| G | 951 | hCoV-19/Brazil/SP-166/2020\|EPI_ISL_523965\|2020-03- | .................... | .................... | .................... | .................... | .................... | .................... |
| G | 952 | hCoV-19/Moldova/ChVir7777/2020\|EPI_ISL_523966\|2020 | .................... | .................... | .................... | .................... | .................... | .................... |
| G | 953 | hCoV-19/Brazil/SP-168/2020\|EPI_ISL_523967\|2020-04- | .................... | .................... | .................... | .................... | .................... | .................... |
| G | 954 | hCoV-19/Moldova/ChVir7776/2020\|EPI_ISL_523968\|2020 | .................... | .................... | .................... | .................... | .................... | .................... |
| G | 955 | hCoV-19/Brazil/SP-170/2020\|EPI_ISL_523969\|2020-03- | .................... | .................... | .................... | .................... | .................... | .................... |
| G | 956 | hCoV-19/Brazil/SP-171/2020\|EPI_ISL_523970\|2020-04- | .................... | .................... | .................... | .................... | .................... | .................... |
| G | 957 | hCoV-19/Brazil/SP-174/2020\|EPI_ISL_523971\|2020-03- | .................... | .................... | .................... | .................... | .................... | .................... |
| G | 958 | hCoV-19/Brazil/SP-176/2020\|EPI_ISL_523972\|2020-04- | .................... | .................... | .................... | .................... | .................... | .................... |
| G | 959 | hCoV-19/Brazil/SP-177/2020\|EPI_ISL_523973\|2020-04- | .................... | .................... | .................... | .................... | .................... | .................... |
| G | 960 | hCoV-19/Brazil/SP-178/2020\|EPI_ISL_523974\|2020-04- | .................... | .................... | .................... | .................... | .................... | .................... |
| G | 961 | hCoV-19/Brazil/SP-179/2020\|EPI_ISL_523975\|2020-04- | .................... | .................... | .................... | .................... | .................... | .................... |
| G | 962 | hCoV-19/Brazil/SP-181/2020\|EPI_ISL_523976\|2020-04- | .................... | .................... | .................... | .................... | .................... | .................... |
| G | 963 | hCoV-19/Brazil/SP-182/2020\|EPI_ISL_523977\|2020-04- | .................... | .................... | .................... | .................... | .................... | .................... |
| G | 964 | hCoV-19/Brazil/SP-195/2020\|EPI_ISL_523978\|2020-04- | .................... | .................... | .................... | .................... | .................... | .................... |
| G | 965 | hCoV-19/Peru/ARE-INS-072/2020\|EPI_ISL_523979\|2020- | .................... | .................... | .................... | .................... | .................... | .................... |
| G | 966 | hCoV-19/Brazil/SP-199/2020\|EPI_ISL_523980\|2020-04- | .................... | .................... | .................... | .................... | .................... | .................... |
| G | 967 | hCoV-19/Brazil/SP-200/2020\|EPI_ISL_523981\|2020-04- | .................... | .................... | .................... | .................... | .................... | .................... |
| G | 968 | hCoV-19/Brazil/SP-190/2020\|EPI_ISL_523982\|2020-03- | .................... | .................... | .................... | .................... | .................... | .................... |
| G | 969 | hCoV-19/Brazil/SP-191/2020\|EPI_ISL_523983\|2020-04- | .................... | .................... | .................... | .................... | .................... | .................... |
| G | 970 | hCoV-19/Brazil/SP-193/2020\|EPI_ISL_523984\|2020-04- | .................... | .................... | .................... | .................... | .................... | .................... |
| G | 971 | hCoV-19/Brazil/SP-202/2020\|EPI_ISL_523985\|2020-04- | .................... | .................... | .................... | .................... | .................... | .................... |
| G | 972 | hCoV-19/Brazil/SP-203/2020\|EPI_ISL_523986\|2020-04- | .................... | .................... | .................... | .................... | .................... | .................... |
| G | 973 | hCoV-19/Brazil/SP-205/2020\|EPI_ISL_523988\|2020-04- | .................... | .................... | .................... | .................... | .................... | .................... |
| G | 974 | hCoV-19/Brazil/SP-208/2020\|EPI_ISL_523989\|2020-04- | .................... | .................... | .................... | .................... | .................... | .................... |
| G | 975 | hCoV-19/Brazil/SP-209/2020\|EPI_ISL_523990\|2020-04- | .................... | .................... | .................... | .................... | .................... | .................... |
| G | 976 | hCoV-19/Brazil/SP-211/2020\|EPI_ISL_523991\|2020-04- | .................... | .................... | .................... | .................... | .................... | .................... |
| G | 977 | hCoV-19/Brazil/SP-213/2020\|EPI_ISL_523992\|2020-04- | .................... | .................... | .................... | .................... | .................... | .................... |
| G | 978 | hCoV-19/Brazil/SP-217/2020\|EPI_ISL_523993\|2020-04- | .................... | .................... | .................... | .................... | .................... | .................... |
| G | 979 | hCoV-19/Peru/LAM-INS-073/2020\|EPI_ISL_523994\|2020- | .................... | .................... | .................... | .................... | .................... | .................... |
| G | 980 | hCoV-19/Russia/SPE-RII-10152V/2020\|EPI_ISL_523995\| | .................... | .................... | .................... | .................... | .................... | .................... |
| G | 981 | hCoV-19/Russia/SPE-RII-10167V/2020\|EPI_ISL_523996\| | .................... | .................... | .................... | .................... | .................... | .................... |
| G | 982 | hCoV-19/Russia/SPE-RII-10325V/2020\|EPI_ISL_523997\| | .................... | .................... | .................... | .................... | .................... | .................... |
| G | 983 | hCoV-19/Russia/SPE-RII-11036V/2020\|EPI_ISL_523998\| | .................... | .................... | .................... | .................... | .................... | .................... |
| G | 984 | hCoV-19/Russia/SPE-RII-11688V/2020\|EPI_ISL_523999\| | .................... | .................... | .................... | .................... | .................... | .................... |
| G | 985 | hCoV-19/India/AP-CCMB-GC41/2020\|EPI_ISL_528809\|202 | .................... | .................... | .................... | .................... | .................... | .................... |
| G | 986 | hCoV-19/India/TG-CCMB-GC1/2020\|EPI_ISL_528810\|2020 | .................... | .................... | .................... | .................... | .................... | .................... |
| G | 987 | hCoV-19/India/TG-CCMB-GC16/2020\|EPI_ISL_528811\|202 | .................... | .................... | .................... | .................... | .................... | .................... |
| G | 988 | hCoV-19/India/TG-CCMB-GC2/2020\|EPI_ISL_528812\|2020 | .................... | .................... | .................... | .................... | .................... | .................... |
| G | 989 | hCoV-19/India/TG-CCMB-GC20/2020\|EPI_ISL_528813\|202 | .................... | .................... | .................... | .................... | .................... | .................... |
| G | 990 | hCoV-19/India/TG-CCMB-GC22/2020\|EPI_ISL_528814\|202 | .................... | .................... | .................... | .................... | .................... | .................... |
| G | 991 | hCoV-19/India/TG-CCMB-GC28/2020\|EPI_ISL_528815\|202 | .................... | .................... | .................... | .................... | .................... | .................... |
| G | 992 | hCoV-19/India/TG-CCMB-GC3/2020\|EPI_ISL_528816\|2020 | .................... | .................... | .................... | .................... | .................... | .................... |
| G | 993 | hCoV-19/India/TG-CCMB-GC39/2020\|EPI_ISL_528817\|202 | .................... | .................... | .................... | .................... | .................... | .................... |
| G | 994 | hCoV-19/India/TG-CCMB-GC4/2020\|EPI_ISL_528818\|2020 | .................... | .................... | .................... | .................... | .................... | .................... |
| G | 995 | hCoV-19/India/TG-CCMB-GC42/2020\|EPI_ISL_528819\|202 | .................... | .................... | .................... | .................... | .................... | .................... |
| G | 996 | hCoV-19/India/TG-CCMB-GC43/2020\|EPI_ISL_528820\|202 | .................... | .................... | .................... | .................... | .................... | .................... |
| G | 997 | hCoV-19/India/TG-CCMB-GC8/2020\|EPI_ISL_528821\|2020 | .................... | .................... | .................... | .................... | .................... | .................... |
| G | 998 | hCoV-19/India/TG-CCMB-GC9/2020\|EPI_ISL_528822\|2020 | .................... | .................... | .................... | .................... | .................... | .................... |
| G | 999 | hCoV-19/India/TG-CCMB-W842/2020\|EPI_ISL_528823\|202 | .................... | .................... | .................... | .................... | .................... | .................... |
| G | 1000 | hCoV-19/India/TG-CCMB-W848/2020\|EPI_ISL_528824\|202 | .................... | .................... | .................... | .................... | .................... | .................... |
| G | 1001 | hCoV-19/India/TG-CCMB-W849/2020\|EPI_ISL_528825\|202 | .................... | .................... | .................... | .................... | .................... | .................... |
| G | 1002 | hCoV-19/India/TG-CCMB-W850/2020\|EPI_ISL_528826\|202 | .................... | .................... | .................... | .................... | .................... | .................... |
| G | 1003 | hCoV-19/India/TG-CCMB-W855/2020\|EPI_ISL_528827\|202 | .................... | .................... | .................... | .................... | .................... | .................... |
| G | 1004 | hCoV-19/India/TG-CCMB-W863/2020\|EPI_ISL_528828\|202 | .................... | .................... | .................... | .................... | .................... | .................... |
| G | 1005 | hCoV-19/India/TG-CCMB-W866/2020\|EPI_ISL_528829\|202 | .................... | .................... | .................... | .................... | .................... | .................... |
| G | 1006 | hCoV-19/India/TG-CCMB-W887/2020\|EPI_ISL_528830\|202 | .................... | .................... | .................... | .................... | .................... | .................... |
| G | 1007 | hCoV-19/India/TG-CCMB-W897/2020\|EPI_ISL_528831\|202 | .................... | .................... | .................... | .................... | .................... | .................... |
| G | 1008 | hCoV-19/India/TG-CCMB-W899/2020\|EPI_ISL_528832\|202 | .................... | .................... | .................... | .................... | .................... | .................... |
| G | 1009 | hCoV-19/India/TG-CCMB-W900/2020\|EPI_ISL_528833\|202 | .................... | .................... | .................... | .................... | .................... | .................... |
| G | 1010 | hCoV-19/India/TG-CCMB-W903/2020\|EPI_ISL_528834\|202 | .................... | .................... | .................... | .................... | .................... | .................... |
| G | 1011 | hCoV-19/India/TG-CCMB-W908/2020\|EPI_ISL_528835\|202 | .................... | .................... | .................... | .................... | .................... | .................... |
| G | 1012 | hCoV-19/India/TG-CCMB-W910/2020\|EPI_ISL_528836\|202 | .................... | .................... | .................... | .................... | .................... | .................... |
| G | 1013 | hCoV-19/India/TG-CCMB-W919/2020\|EPI_ISL_528837\|202 | .................... | .................... | .................... | .................... | .................... | .................... |
| G | 1014 | hCoV-19/India/TG-CCMB-W990/2020\|EPI_ISL_528838\|202 | .................... | .................... | .................... | .................... | .................... | .................... |
| G | 1015 | hCoV-19/India/TG-CCMB-W996/2020\|EPI_ISL_528839\|202 | .................... | .................... | .................... | .................... | .................... | .................... |
| G | 1016 | hCoV-19/India/TG-CCMB-W998/2020\|EPI_ISL_528840\|202 | .................... | .................... | .................... | .................... | .................... | .................... |
| G | 1017 | hCoV-19/India/TG-CCMB-X10/2020\|EPI_ISL_528841\|2020 | .................... | .................... | .................... | .................... | .................... | .................... |
| G | 1018 | hCoV-19/India/TG-CCMB-X100/2020\|EPI_ISL_528842\|202 | .................... | .................... | .................... | .................... | .................... | .................... |
| G | 1019 | hCoV-19/India/TG-CCMB-X102/2020\|EPI_ISL_528843\|202 | .................... | .................... | .................... | .................... | .................... | .................... |
| G | 1020 | hCoV-19/India/TG-CCMB-X103/2020\|EPI_ISL_528844\|202 | .................... | .................... | .................... | .................... | .................... | .................... |
| G | 1021 | hCoV-19/India/TG-CCMB-X110/2020\|EPI_ISL_528845\|202 | .................... | .................... | .................... | .................... | .................... | .................... |
| G | 1022 | hCoV-19/India/TG-CCMB-X111/2020\|EPI_ISL_528846\|202 | .................... | .................... | .................... | .................... | .................... | .................... |
| G | 1023 | hCoV-19/India/TG-CCMB-X112/2020\|EPI_ISL_528847\|202 | .................... | .................... | .................... | .................... | .................... | .................... |

```
G 1024 hCoV-19/India/TG-CCMB-X113/2020|EPI_ISL_528848|202 ···················· ···················· ···················· ···················· ···················· ····················
G 1025 hCoV-19/Indonesia/YO-UGM-202449/2020|EPI_ISL_51680 ···················· ···················· ···················· ···················· ···················· ····················
G 1026 hCoV-19/Singapore/829/2020|EPI_ISL_516801|2020-03- ···················· ···················· ···················· ···················· ···················· ····················
G 1027 hCoV-19/Singapore/830/2020|EPI_ISL_516802|2020-03- ···················· ···················· ···················· ···················· ···················· ····················
G 1028 hCoV-19/Singapore/831/2020|EPI_ISL_516803|2020-03- ···················· ···················· ···················· ···················· ···················· ····················
G 1029 hCoV-19/Singapore/832/2020|EPI_ISL_516804|2020-03- ···················· ···················· ···················· ···················· ···················· ····················
G 1030 hCoV-19/Singapore/833/2020|EPI_ISL_516805|2020-03- ···················· ···················· ···················· ···················· ···················· ····················
G 1031 hCoV-19/Indonesia/YO-UGM-200927/2020|EPI_ISL_51680 ···················· ···················· ···················· ···················· ···················· ····················
G 1032 hCoV-19/Singapore/807/2020|EPI_ISL_516807|2020-04- ···················· ···················· ···················· ···················· ···················· ····················
G 1033 hCoV-19/Singapore/808/2020|EPI_ISL_516808|2020-08- ···················· ···················· ···················· ···················· ···················· ····················
G 1034 hCoV-19/Singapore/809/2020|EPI_ISL_516809|2020-07- ···················· ···················· ···················· ···················· ···················· ····················
G 1035 hCoV-19/Singapore/810/2020|EPI_ISL_516810|2020-08- ···················· A··················· ···················· ···················· ···················· ····················
G 1036 hCoV-19/Singapore/811/2020|EPI_ISL_516811|2020-08- ···················· ···················· ···················· ···················· ···················· ····················
G 1037 hCoV-19/Singapore/812/2020|EPI_ISL_516812|2020-08- ···················· ···················· ···················· ···················· ···················· ····················
G 1038 hCoV-19/Singapore/813/2020|EPI_ISL_516813|2020-08- ···················· ···················· ···················· ···················· ···················· ····················
G 1039 hCoV-19/Singapore/814/2020|EPI_ISL_516814|2020-08- ···················· ···················· ···················· ···················· ···················· ····················
G 1040 hCoV-19/Singapore/815/2020|EPI_ISL_516815|2020-08- ···················· ···················· ···················· ···················· ···················· ····················
G 1041 hCoV-19/Singapore/816/2020|EPI_ISL_516816|2020-08- ···················· ···················· ···················· ···················· ···················· ····················
G 1042 hCoV-19/Singapore/817/2020|EPI_ISL_516817|2020-08- ···················· ···················· ···················· ···················· ···················· ····················
G 1043 hCoV-19/Singapore/818/2020|EPI_ISL_516818|2020-07- ···················· ···················· ···················· ···················· ···················· ····················
G 1044 hCoV-19/Singapore/819/2020|EPI_ISL_516819|2020-08- ···················· ···················· ···················· ···················· ···················· ····················
G 1045 hCoV-19/Singapore/820/2020|EPI_ISL_516820|2020-08- ···················· ···················· ···················· ···················· ···················· ····················
G 1046 hCoV-19/Singapore/821/2020|EPI_ISL_516821|2020-07- ···················· A··················· ···················· ···················· ···················· ····················
G 1047 hCoV-19/Singapore/822/2020|EPI_ISL_516822|2020-07- ···················· ···················· ···················· ···················· ···················· ····················
G 1048 hCoV-19/Singapore/823/2020|EPI_ISL_516823|2020-08- ···················· ·A·················· ···················· ···················· ···················· ··A·················
G 1049 hCoV-19/Singapore/824/2020|EPI_ISL_516824|2020-08- ···················· ···················· ···················· ···················· ···················· ····················
G 1050 hCoV-19/Singapore/825/2020|EPI_ISL_516825|2020-08- ···················· ···················· ···················· ···················· ···················· ····················
G 1051 hCoV-19/England/LOND-125FCEB/2020|EPI_ISL_516831|2 ···················· ···················· ···················· ···················· ···················· ····················
G 1052 hCoV-19/England/LOND-1260986/2020|EPI_ISL_516832|2 ···················· ···················· ···················· ···················· ···················· ····················
G 1053 hCoV-19/England/LOND-1260713/2020|EPI_ISL_516833|2 ···················· ···················· ···················· ···················· ···················· ····················
G 1054 hCoV-19/England/LOND-12608F2/2020|EPI_ISL_516834|2 ···················· ···················· ···················· ···················· ···················· ····················
G 1055 hCoV-19/England/LOND-12606CB/2020|EPI_ISL_516835|2 ···················· ···················· ···················· ···················· ···················· ····················
G 1056 hCoV-19/England/LOND-125FC63/2020|EPI_ISL_516836|2 ···T················ ···················· ···················· ···················· ···················· ····················
G 1057 hCoV-19/England/LOND-125FA96/2020|EPI_ISL_516880|2 ···················· ···················· ···················· ···················· ···················· ····················
G 1058 hCoV-19/England/LOND-125FAA5/2020|EPI_ISL_516883|2 ···················· ···················· ···················· ···················· ···················· ····················
G 1059 hCoV-19/Israel/CVL-n17112/2020|EPI_ISL_516886|2020 ···················· ···················· ···················· ···················· ···················· ····················
G 1060 hCoV-19/Israel/CVL-n17372/2020|EPI_ISL_516887|2020 ···················· ···················· ···················· ···················· ···················· ····················
G 1061 hCoV-19/Israel/CVL-n17884/2020|EPI_ISL_516888|2020 ···················· ···················· ···················· ···················· ···················· ····················
G 1062 hCoV-19/Israel/CVL-n17916/2020|EPI_ISL_516889|2020 ···················· ···················· ···················· ···················· ···················· ····················
G 1063 hCoV-19/Israel/CVL-n17925/2020|EPI_ISL_516890|2020 ···················· ···················· ···················· ···················· ···················· ····················
G 1064 hCoV-19/Israel/CVL-n17953/2020|EPI_ISL_516891|2020 ···················· ···················· ···················· ···················· ···················· ····················
G 1065 hCoV-19/Israel/CVL-n18311/2020|EPI_ISL_516892|2020 ···················· ···················· ···················· ···················· ···················· ····················
G 1066 hCoV-19/Israel/CVL-n18346/2020|EPI_ISL_516893|2020 ···················· ···················· ···················· ···················· ···················· ····················
G 1067 hCoV-19/Israel/CVL-n18352/2020|EPI_ISL_516894|2020 ···················· ···················· ···················· ···················· ···················· ····················
G 1068 hCoV-19/Israel/CVL-n18416/2020|EPI_ISL_516895|2020 ···················· ···················· ···················· ···················· ···················· ····················
G 1069 hCoV-19/Israel/CVL-n18467/2020|EPI_ISL_516896|2020 ···················· ···················· ···················· ···················· ···················· ····················
G 1070 hCoV-19/Israel/CVL-n19018/2020|EPI_ISL_516897|2020 ···················· ···················· ···················· ···················· ···················· ····················
G 1071 hCoV-19/Israel/CVL-n19042/2020|EPI_ISL_516898|2020 ···················· ···················· ···················· ···················· ···················· ····················
G 1072 hCoV-19/Israel/CVL-n19050/2020|EPI_ISL_516899|2020 ···················· ···················· ···················· ···················· ···················· ····················
G 1073 hCoV-19/Australia/VIC4672/2020|EPI_ISL_519060|2020 ···················· ···················· ···················· ···················· ···················· ····················
G 1074 hCoV-19/Australia/VIC4673/2020|EPI_ISL_519061|2020 ···················· ···················· ···················· ···················· ···················· ····················
G 1075 hCoV-19/Australia/VIC4674/2020|EPI_ISL_519062|2020 ···················· ···················· ···················· ···················· ···················· ····················
G 1076 hCoV-19/Australia/VIC4675/2020|EPI_ISL_519063|2020 ···················· ···················· ···················· ···················· ···················· ····················
G 1077 hCoV-19/Australia/VIC4676/2020|EPI_ISL_519064|2020 ···················· ···················· ···················· ···················· ···················· ····················
G 1078 hCoV-19/Australia/VIC4677/2020|EPI_ISL_519065|2020 ···················· ···················· ···················· ···················· ···················· ····················
G 1079 hCoV-19/Australia/VIC4678/2020|EPI_ISL_519066|2020 ···················· ···················· ···················· ···················· ···················· ····················
G 1080 hCoV-19/Australia/VIC4679/2020|EPI_ISL_519067|2020 ···················· ···················· ···················· ···················· ···················· ····················
G 1081 hCoV-19/Australia/VIC4680/2020|EPI_ISL_519068|2020 ···················· ···················· ···················· ···················· ···················· ····················
G 1082 hCoV-19/Australia/VIC4681/2020|EPI_ISL_519069|2020 ···················· ···················· ···················· ···················· ···················· ····················
G 1083 hCoV-19/Australia/VIC4682/2020|EPI_ISL_519070|2020 ···················· ···················· ···················· ···················· ···················· ····················
G 1084 hCoV-19/Australia/VIC4683/2020|EPI_ISL_519071|2020 ···················· ···················· ···················· ···················· ···················· ····················
G 1085 hCoV-19/Australia/VIC4684/2020|EPI_ISL_519072|2020 ···················· ···················· ···················· ···················· ···················· ····················
G 1086 hCoV-19/Australia/VIC4685/2020|EPI_ISL_519073|2020 ···················· ···················· ···················· ···················· ···················· ····················
G 1087 hCoV-19/Australia/VIC4686/2020|EPI_ISL_519074|2020 ···················· ···················· ···················· ···················· ···················· ····················
G 1088 hCoV-19/Australia/VIC4687/2020|EPI_ISL_519075|2020 ···················· ···················· ···················· ···················· ···················· ····················
G 1089 hCoV-19/Australia/VIC4688/2020|EPI_ISL_519076|2020 ···················· ···················· ···················· ···················· ···················· ····················
G 1090 hCoV-19/Australia/VIC4689/2020|EPI_ISL_519077|2020 ···················· ···················· ···················· ···················· ···················· ····················
G 1091 hCoV-19/Australia/VIC4690/2020|EPI_ISL_519078|2020 ···················· ···················· ···················· ···················· ···················· ····················
G 1092 hCoV-19/Australia/VIC4691/2020|EPI_ISL_519079|2020 ···················· ···················· ···················· ···················· ···················· ····················
G 1093 hCoV-19/Australia/VIC4692/2020|EPI_ISL_519080|2020 ···················· ···················· ···················· ···················· ···················· ····················
G 1094 hCoV-19/Australia/VIC4693/2020|EPI_ISL_519081|2020 ···················· ···················· ···················· ···················· ···················· ····················
G 1095 hCoV-19/Australia/VIC4694/2020|EPI_ISL_519082|2020 ···················· ···················· ···················· ···················· ···················· ····················
G 1096 hCoV-19/Australia/VIC4695/2020|EPI_ISL_519083|2020 ···················· ···················· ···················· ···················· ···················· ····················
G 1097 hCoV-19/Australia/VIC4696/2020|EPI_ISL_519084|2020 ···················· ···················· ···················· ···················· ···················· ····················
G 1098 hCoV-19/Australia/VIC4697/2020|EPI_ISL_519085|2020 ···················· ···················· ···················· ···················· ···················· ····················
G 1099 hCoV-19/Australia/VIC4698/2020|EPI_ISL_519086|2020 ···················· ···················· ···················· ···················· ···················· ····················
G 1100 hCoV-19/Australia/VIC4699/2020|EPI_ISL_519087|2020 ···················· ···················· ···················· ···················· ···················· ····················
G 1101 hCoV-19/Australia/VIC4700/2020|EPI_ISL_519088|2020 ···················· ···················· ···················· ···················· ···················· ····················
G 1102 hCoV-19/Australia/VIC4701/2020|EPI_ISL_519089|2020 ···················· ···················· ···················· ···················· ···················· ····················
G 1103 hCoV-19/Australia/VIC4702/2020|EPI_ISL_519090|2020 ···················· ···················· ···················· ···················· ···················· ····················
```

G 1104 hCoV-19/Australia/VIC4703/2020|EPI_ISL_519091|2020 .................... .................... ..................... .................... .................... ....................
G 1105 hCoV-19/Australia/VIC4704/2020|EPI_ISL_519092|2020 .................... .................... ..................... .................... .................... ....................
G 1106 hCoV-19/Australia/VIC4705/2020|EPI_ISL_519093|2020 .................... .................... ..................... .................... .................... ....................
G 1107 hCoV-19/Australia/VIC4706/2020|EPI_ISL_519094|2020 .................... .................... ..................... .................... .................... ....................
G 1108 hCoV-19/Australia/VIC4707/2020|EPI_ISL_519095|2020 .................... .................... ..................... .................... .................... ....................
G 1109 hCoV-19/Australia/VIC4708/2020|EPI_ISL_519096|2020 .................... .................... ..................... .................... .................... ....................
G 1110 hCoV-19/Australia/VIC4709/2020|EPI_ISL_519097|2020 .................... .................... ..................... .................... .................... ....................
G 1111 hCoV-19/Australia/VIC4710/2020|EPI_ISL_519098|2020 .................... .................... ..................... .................... .................... ....................
G 1112 hCoV-19/Australia/VIC4711/2020|EPI_ISL_519099|2020 .................... .................... ..................... .................... .................... ....................
G 1113 hCoV-19/Australia/VIC5285/2020|EPI_ISL_520049|2020 .................... .................... ..................... .................... .................... ....................
G 1114 hCoV-19/Australia/VIC5286/2020|EPI_ISL_520050|2020 .................... .................... ..................... .................... .................... ....................
G 1115 hCoV-19/Australia/VIC5289/2020|EPI_ISL_520052|2020 .................... .................... ..................... .................... .................... ....................
G 1116 hCoV-19/Australia/VIC5290/2020|EPI_ISL_520053|2020 .................... .................... ..................... .................... .................... ....................
G 1117 hCoV-19/Australia/VIC5291/2020|EPI_ISL_520054|2020 .................... .................... ..................... .................... .................... ....................
G 1118 hCoV-19/Australia/VIC5292/2020|EPI_ISL_520055|2020 .................... .................... ..................... .................... .................... ....................
G 1119 hCoV-19/Australia/VIC5293/2020|EPI_ISL_520056|2020 .................... .................... ..................... .................... .................... ....................
G 1120 hCoV-19/Australia/VIC5294/2020|EPI_ISL_520057|2020 .................... .................... ..................... .................... .................... ....................
G 1121 hCoV-19/Australia/VIC5295/2020|EPI_ISL_520058|2020 .................... .................... ..................... .................... .................... ....................
G 1122 hCoV-19/Australia/VIC5296/2020|EPI_ISL_520059|2020 .................... .................... ..................... .................... .................... ....................
G 1123 hCoV-19/Australia/VIC5297/2020|EPI_ISL_520060|2020 .................... .................... ..................... .................... .................... ....................
G 1124 hCoV-19/Australia/VIC5298/2020|EPI_ISL_520061|2020 .................... .................... ..................... .................... .................... ....................
G 1125 hCoV-19/Australia/VIC5299/2020|EPI_ISL_520062|2020 .................... .................... ..................... .................... .................... ....................
G 1126 hCoV-19/Australia/VIC5300/2020|EPI_ISL_520063|2020 .................... .................... ..................... .................... .................... ....................
G 1127 hCoV-19/Australia/VIC5302/2020|EPI_ISL_520065|2020 .................... .................... ..................... .................... .................... ....................
G 1128 hCoV-19/Australia/VIC5303/2020|EPI_ISL_520066|2020 .................... .................... ..................... .................... .................... ....................
G 1129 hCoV-19/Australia/VIC5304/2020|EPI_ISL_520067|2020 .................... .................... ..................... .................... .................... ....................
G 1130 hCoV-19/Australia/VIC5305/2020|EPI_ISL_520068|2020 .................... .................... ..................... .................... .................... ....................
G 1131 hCoV-19/Australia/VIC5306/2020|EPI_ISL_520069|2020 .................... .................... ..................... .................... .................... ....................
G 1132 hCoV-19/Australia/VIC5307/2020|EPI_ISL_520070|2020 .................... .................... ..................... .................... .................... ....................
G 1133 hCoV-19/Australia/VIC5308/2020|EPI_ISL_520071|2020 .................... .................... ..................... .................... .................... ....................
G 1134 hCoV-19/Australia/VIC5309/2020|EPI_ISL_520072|2020 .................... .................... ..................... .................... .................... ....................
G 1135 hCoV-19/Australia/VIC5310/2020|EPI_ISL_520073|2020 .................... .................... ..................... .................... .................... ....................
G 1136 hCoV-19/Australia/VIC5311/2020|EPI_ISL_520074|2020 .................... .................... ..................... .................... .................... ....................
G 1137 hCoV-19/Australia/VIC5313/2020|EPI_ISL_520076|2020 .................... .................... ..................... .................... .................... ....................
G 1138 hCoV-19/Australia/VIC5314/2020|EPI_ISL_520077|2020 .................... .................... ..................... .................... .................... ....................
G 1139 hCoV-19/Australia/VIC5315/2020|EPI_ISL_520078|2020 .................... .................... ..................... .................... .................... ....................
G 1140 hCoV-19/Australia/VIC5317/2020|EPI_ISL_520080|2020 .................... .................... ..................... .................... .................... ....................
G 1141 hCoV-19/Australia/VIC5318/2020|EPI_ISL_520081|2020 .................... .................... ..................... .................... .................... ....................
G 1142 hCoV-19/Australia/VIC5319/2020|EPI_ISL_520082|2020 .................... .................... ..................... .................... .................... ....................
G 1143 hCoV-19/Australia/VIC5320/2020|EPI_ISL_520083|2020 .................... .................... ..................... .................... .................... ....................
G 1144 hCoV-19/Australia/VIC5321/2020|EPI_ISL_520084|2020 .................... .................... ..................... .................... .................... ....................
G 1145 hCoV-19/Australia/VIC5322/2020|EPI_ISL_520085|2020 .................... .................... ..................... .................... .................... ....................
G 1146 hCoV-19/Australia/VIC5323/2020|EPI_ISL_520086|2020 .................... .................... ..................... .................... .................... ....................
G 1147 hCoV-19/Australia/VIC5324/2020|EPI_ISL_520087|2020 .................... .................... ..................... .................... .................... ....................
G 1148 hCoV-19/Australia/VIC5325/2020|EPI_ISL_520088|2020 .................... .................... ..................... .................... .................... ....................
G 1149 hCoV-19/Australia/VIC5326/2020|EPI_ISL_520089|2020 .................... .................... ..................... .................... .................... ....................
G 1150 hCoV-19/Australia/VIC6618/2020|EPI_ISL_521380|2020 .................... .................... ..................... .................... .................... ....................
G 1151 hCoV-19/Australia/VIC6619/2020|EPI_ISL_521381|2020 .................... .................... ..................... .................... .................... ....................
G 1152 hCoV-19/Australia/VIC6620/2020|EPI_ISL_521382|2020 .................... .................... ..................... .................... .................... ....................
G 1153 hCoV-19/Australia/VIC6622/2020|EPI_ISL_521383|2020 .................... .................... ..................... .................... .................... ....................
G 1154 hCoV-19/Australia/VIC6624/2020|EPI_ISL_521384|2020 .................... .................... ..................... .................... .................... ....................
G 1155 hCoV-19/Australia/VIC6625/2020|EPI_ISL_521385|2020 .................... .................... ..................... .................... .................... ....................
G 1156 hCoV-19/Australia/VIC6626/2020|EPI_ISL_521386|2020 .................... .................... ..................... .................... .................... ....................
G 1157 hCoV-19/Australia/VIC6627/2020|EPI_ISL_521387|2020 .................... .................... ..................... .................... .................... ....................
G 1158 hCoV-19/Australia/VIC6628/2020|EPI_ISL_521388|2020 .................... .................... ..................... .................... .................... ....................
G 1159 hCoV-19/Australia/VIC6629/2020|EPI_ISL_521389|2020 .................... .................... ..................... .................... .................... ....................
G 1160 hCoV-19/Australia/VIC6631/2020|EPI_ISL_521391|2020 .................... .................... ..................... .................... .................... ....................
G 1161 hCoV-19/Australia/VIC6634/2020|EPI_ISL_521394|2020 .................... .................... ..................... .................... .................... ....................
G 1162 hCoV-19/Australia/VIC6636/2020|EPI_ISL_521395|2020 .................... .................... ..................... .................... .................... ....................
G 1163 hCoV-19/Australia/VIC6637/2020|EPI_ISL_521396|2020 .................... .................... ..................... .................... .................... ....................
G 1164 hCoV-19/Australia/VIC6639/2020|EPI_ISL_521398|2020 .................... .................... ..................... .................... .................... ....................
G 1165 hCoV-19/Australia/VIC6640/2020|EPI_ISL_521399|2020 .................... .................... ..................... .................... .................... ....................
G 1166 hCoV-19/Australia/NSW858/2020|EPI_ISL_526200|2020- .................... .................... ..................... .................... .................... ....................
G 1167 hCoV-19/Australia/NSW859/2020|EPI_ISL_526201|2020- .................... .................... ..................... .................... .................... ....................
G 1168 hCoV-19/Australia/NSW860/2020|EPI_ISL_526202|2020- .................... .................... ..................... .................... .................... ....................
G 1169 hCoV-19/Australia/NSW861/2020|EPI_ISL_526203|2020- .................... .................... ..................... .................... .................... ....................
G 1170 hCoV-19/Australia/NSW862/2020|EPI_ISL_526204|2020- .................... .................... ..................... .................... .................... ....................
G 1171 hCoV-19/Australia/NSW863/2020|EPI_ISL_526205|2020- .................... .................... ..................... .................... .................... ....................
G 1172 hCoV-19/Australia/NSW864/2020|EPI_ISL_526206|2020- .................... .................... ..................... .................... .................... ....................
G 1173 hCoV-19/Australia/NSW865/2020|EPI_ISL_526207|2020- .................... .................... ..................... .................... .................... ....................
G 1174 hCoV-19/Australia/NSW866/2020|EPI_ISL_526208|2020- .................... .................... ..................... .................... .................... ....................
G 1175 hCoV-19/Australia/NSW867/2020|EPI_ISL_526209|2020- .................... .................... ..................... .................... .................... ....................
G 1176 hCoV-19/Australia/NSW868/2020|EPI_ISL_526210|2020- .................... .................... ..................... .................... .................... ....................
G 1177 hCoV-19/Australia/NSW869/2020|EPI_ISL_526211|2020- .................... .................... ..................... .................... .................... ....................
G 1178 hCoV-19/Australia/NSW870/2020|EPI_ISL_526212|2020- .................... .................... ..................... .................... .................... ....................
G 1179 hCoV-19/Australia/NSW871/2020|EPI_ISL_526213|2020- .................... .................... ..................... .................... .................... ....................
G 1180 hCoV-19/Australia/NSW872/2020|EPI_ISL_526214|2020- .................... .................... ..................... .................... .................... ....................
G 1181 hCoV-19/Hungary/MH-597/2020|EPI_ISL_526215|2020-04 .................... .................... ..................... .................... .................... ....................
G 1182 hCoV-19/Hungary/MH-642/2020|EPI_ISL_526216|2020-04 .................... .................... ..................... .................... .................... ....................
G 1183 hCoV-19/Hungary/MH-732/2020|EPI_ISL_526217|2020-04 .................... .................... ..................... .................... .................... ....................

G 1184 hCoV-19/Hungary/MH-1403/2020|EPI_ISL_526218|2020-0 ···················· ···················· ···················· ···················· ···················· ····················
G 1185 hCoV-19/Hungary/MH-6074/2020|EPI_ISL_526219|2020-0 ···················· ·A·················· ···················· ···················· ···················· ····················
G 1186 hCoV-19/Hungary/MH-6633/2020|EPI_ISL_526220|2020-0 ···················· ···················· ···················· ···················· ···················· ····················
G 1187 hCoV-19/Hungary/MH-6657/2020|EPI_ISL_526221|2020-0 ···················· ···················· ···················· ···················· ···················· ····················
G 1188 hCoV-19/Hungary/MH-7051/2020|EPI_ISL_526222|2020-0 ···················· ···················· ···················· ···················· ···················· ····················
G 1189 hCoV-19/Hungary/MH-8268/2020|EPI_ISL_526223|2020-0 ···················· ···················· ···················· ···················· ···················· ····················
G 1190 hCoV-19/Hungary/MH-9653/2020|EPI_ISL_526224|2020-0 ···················· ···················· ···················· ···················· ···················· ····················
G 1191 hCoV-19/Hungary/MH-13211/2020|EPI_ISL_526225|2020- ···················· ·A·················· ···················· ···················· ···················· ····················
G 1192 hCoV-19/Hungary/MH-1421/2020|EPI_ISL_526226|2020-0 ···················· ···················· ···················· ···················· ···················· ····················
G 1193 hCoV-19/Hungary/MH-5846/2020|EPI_ISL_526227|2020-0 ···················· ···················· ···················· ···················· ···················· ····················
G 1194 hCoV-19/Hungary/MH-5657/2020|EPI_ISL_526228|2020-0 ···················· ···················· ···················· ···················· ···················· ····················
G 1195 hCoV-19/Hungary/MH-722/2020|EPI_ISL_526229|2020-04 ···················· ···················· ···················· ···················· ···················· ····················
G 1196 hCoV-19/Hungary/MH-879/2020|EPI_ISL_526230|2020-04 ···················· ···················· ···················· ···················· ···················· ····················
G 1197 hCoV-19/Hungary/MH-975/2020|EPI_ISL_526231|2020-04 ···················· ···················· ···················· ···················· ···················· ····················
G 1198 hCoV-19/Hungary/MH-789/2020|EPI_ISL_526232|2020-04 ···················· ···················· ···················· ···················· ···················· ····················
G 1199 hCoV-19/Hungary/MH-824/2020|EPI_ISL_526233|2020-04 ···················· ···················· ···················· ···················· ···················· ····················
G 1200 hCoV-19/Hungary/MH-726/2020|EPI_ISL_526234|2020-04 ···················· ···················· ···················· ···················· ···················· ····················
G 1201 hCoV-19/Hungary/MH-1101/2020|EPI_ISL_526235|2020-0 ···················· ···················· ···················· ···················· ···················· ····················
G 1202 hCoV-19/Hungary/MH-6603/2020|EPI_ISL_526236|2020-0 ···················· ···················· ···················· ···················· ···················· ····················
G 1203 hCoV-19/Hungary/MH-5984/2020|EPI_ISL_526237|2020-0 ···················· ···················· ···················· ···················· ···················· ····················
G 1204 hCoV-19/Hungary/MH-1106/2020|EPI_ISL_526238|2020-0 ···················· ···················· ···················· ···················· ···················· ····················
G 1205 hCoV-19/Canada/ON-UHTC_0104/2020|EPI_ISL_526259|20 ···················· ···················· ···················· ···················· ···················· ····················
G 1206 hCoV-19/Canada/ON-UHTC_0123/2020|EPI_ISL_526268|20 ···················· ···················· ···················· ···················· ···················· ····················
G 1207 hCoV-19/DRC/10041/2020|EPI_ISL_527543|2020-05-22 ···················· ···G················ ···················· ···················· ···················· ····················
G 1208 hCoV-19/DRC/10070/2020|EPI_ISL_527544|2020-05-22 ···················· ···················· ···················· ···················· ···················· ····················
G 1209 hCoV-19/DRC/10324/2020|EPI_ISL_527545|2020-05-22 ···················· ···················· ···················· ···················· ···················· ····················
G 1210 hCoV-19/DRC/10346/2020|EPI_ISL_527546|2020-05-22 ···················· ···················· ···················· ···················· ···················· ····················
G 1211 hCoV-19/DRC/10502/2020|EPI_ISL_527547|2020-05-20 ···················· ···················· ···················· ···················· ···················· ····················
G 1212 hCoV-19/DRC/10533/2020|EPI_ISL_527548|2020-05-20 ···················· ···················· ···················· ···················· ···················· ····················
G 1213 hCoV-19/DRC/10644/2020|EPI_ISL_527549|2020-05-22 ···················· ···················· ···················· ···················· ···················· ····················
G 1214 hCoV-19/DRC/11005/2020|EPI_ISL_527550|2020-05-24 ···················· ···················· ···················· ···················· ···················· ····················
G 1215 hCoV-19/DRC/11022/2020|EPI_ISL_527551|2020-05-24 ···················· ···················· ···················· ···················· ···················· ····················
G 1216 hCoV-19/DRC/11024/2020|EPI_ISL_527552|2020-05-24 ···················· ···················· ···················· ···················· ···················· ····················
G 1217 hCoV-19/DRC/11026/2020|EPI_ISL_527553|2020-05-24 ···················· ···················· ···················· ···················· ···················· ····················
G 1218 hCoV-19/DRC/11031/2020|EPI_ISL_527554|2020-05-24 ···················· ···················· ···················· ···················· ···················· ····················
G 1219 hCoV-19/DRC/11041/2020|EPI_ISL_527555|2020-05-24 ···················· ···················· ···················· ···················· ···················· ····················
G 1220 hCoV-19/DRC/11143/2020|EPI_ISL_527556|2020-05-24 ···················· ···················· ···················· ···················· ···················· ····················
G 1221 hCoV-19/DRC/11252/2020|EPI_ISL_527557|2020-05-21 ···················· ···················· ···················· ···················· ···················· ····················
G 1222 hCoV-19/DRC/11260/2020|EPI_ISL_527558|2020-05-23 ···················· ···················· ···················· ···················· ···················· ····················
G 1223 hCoV-19/DRC/11318/2020|EPI_ISL_527559|2020-05-25 ···················· ···················· ···················· ···················· ···················· ····················
G 1224 hCoV-19/DRC/11348/2020|EPI_ISL_527560|2020-05-25 ···················· ···················· ···················· ···················· ···················· ····················
G 1225 hCoV-19/DRC/11411/2020|EPI_ISL_527561|2020-05-25 ···················· ···················· ···················· ···················· ···················· ····················
G 1226 hCoV-19/DRC/11460/2020|EPI_ISL_527562|2020-05-25 ···················· ···················· ···················· ···················· ···················· ····················
G 1227 hCoV-19/DRC/11477/2020|EPI_ISL_527563|2020-05-25 ···················· ···················· ···················· ···················· ···················· ····················
G 1228 hCoV-19/DRC/11486/2020|EPI_ISL_527564|2020-05-25 ···················· ···················· ···················· ···················· ···················· ····················
G 1229 hCoV-19/DRC/11572/2020|EPI_ISL_527565|2020-05-25 ···················· ···················· ···················· ···················· ···················· ····················
G 1230 hCoV-19/DRC/11623/2020|EPI_ISL_527566|2020-05-25 ···················· ···················· ···················· ···················· ···················· ····················
G 1231 hCoV-19/DRC/11624/2020|EPI_ISL_527567|2020-05-25 ···················· ···················· ···················· ···················· ···················· ····················
G 1232 hCoV-19/DRC/11631/2020|EPI_ISL_527568|2020-05-25 ···················· ···················· ···················· ···················· ···················· ····················
G 1233 hCoV-19/DRC/11648/2020|EPI_ISL_527569|2020-05-25 ···················· ···················· ···················· ···················· ···················· ····················
G 1234 hCoV-19/DRC/11652/2020|EPI_ISL_527570|2020-05-25 ···················· ···················· ···················· ···················· ···················· ····················
G 1235 hCoV-19/DRC/11733/2020|EPI_ISL_527571|2020-05-26 ···················· ···················· ···················· ···················· ···················· ····················
G 1236 hCoV-19/DRC/11806/2020|EPI_ISL_527572|2020-05-26 ···················· ···················· ···················· ···················· ···················· ····················
G 1237 hCoV-19/USA/MN-MDH-1508/2020|EPI_ISL_527573|2020-0 ···················· ···················· ···················· ···················· ···················· ····················
G 1238 hCoV-19/USA/MN-MDH-1509/2020|EPI_ISL_527574|2020-0 ···················· ···················· ···················· ···················· ···················· ····················
G 1239 hCoV-19/USA/MN-MDH-1510/2020|EPI_ISL_527575|2020-0 ···················· ·A·················· ···················· ···················· ···················· ····················
G 1240 hCoV-19/USA/MN-MDH-1511/2020|EPI_ISL_527576|2020-0 ···················· ·A·················· ···················· ···················· ···················· ····················
G 1241 hCoV-19/USA/MN-MDH-1512/2020|EPI_ISL_527577|2020-0 ···················· ·A·················· ···················· ···················· ···················· ····················
G 1242 hCoV-19/USA/MN-MDH-1513/2020|EPI_ISL_527578|2020-0 ···················· ·A·················· ···················· ···················· ···················· ····················
G 1243 hCoV-19/USA/MN-MDH-1514/2020|EPI_ISL_527579|2020-0 ···················· ·A·················· ···················· ···················· ···················· ····················
G 1244 hCoV-19/USA/MN-MDH-1515/2020|EPI_ISL_527580|2020-0 ···················· ·A·················· ···················· ···················· ···················· ····················
G 1245 hCoV-19/USA/MN-MDH-1516/2020|EPI_ISL_527581|2020-0 ···················· ·A·················· ···················· ···················· ···················· ····················
G 1246 hCoV-19/USA/MN-MDH-1517/2020|EPI_ISL_527582|2020-0 ···················· ·A·················· ···················· ···················· ···················· ····················
G 1247 hCoV-19/USA/MN-MDH-1518/2020|EPI_ISL_527583|2020-0 ···················· ·A·················· ···················· ···················· ···················· ····················
G 1248 hCoV-19/USA/MN-MDH-1519/2020|EPI_ISL_527584|2020-0 ···················· ·A·················· ···················· ···················· ···················· ····················
G 1249 hCoV-19/USA/MN-MDH-1520/2020|EPI_ISL_527585|2020-0 ···················· ·A·················· ···················· ···················· ···················· ····················
G 1250 hCoV-19/USA/MN-MDH-1521/2020|EPI_ISL_527586|2020-0 ···················· ·A·················· ···················· ···················· ···················· ····················
G 1251 hCoV-19/USA/MN-MDH-1522/2020|EPI_ISL_527587|2020-0 ···················· ···················· ···················· ···················· ···················· ····················
G 1252 hCoV-19/USA/MN-MDH-1523/2020|EPI_ISL_527588|2020-0 ···················· ·A·················· ···················· ···················· ···················· ····················
G 1253 hCoV-19/USA/MN-MDH-1524/2020|EPI_ISL_527589|2020-0 ···················· ·A·················· ···················· ···················· ···················· ····················
G 1254 hCoV-19/USA/MN-MDH-1525/2020|EPI_ISL_527590|2020-0 ···················· ·A·················· ···················· ···················· ···················· ····················
G 1255 hCoV-19/USA/MN-MDH-1526/2020|EPI_ISL_527591|2020-0 ···················· ·A·················· ···················· ···················· ···················· ····················
G 1256 hCoV-19/USA/MN-MDH-1527/2020|EPI_ISL_527592|2020-0 ···················· ·A·················· ···················· ···················· ···················· ····················
G 1257 hCoV-19/USA/MN-MDH-1528/2020|EPI_ISL_527593|2020-0 ···················· ·A·················· ···················· ···················· ···················· ····················
G 1258 hCoV-19/USA/MN-MDH-1529/2020|EPI_ISL_527594|2020-0 ···················· ·A·················· ···················· ···················· ···················· ····················
G 1259 hCoV-19/USA/MN-MDH-1530/2020|EPI_ISL_527595|2020-0 ···················· ·A·················· ···················· ···················· ···················· ····················
G 1260 hCoV-19/USA/MN-MDH-1531/2020|EPI_ISL_527596|2020-0 ···················· ·A·················· ···················· ···················· ···················· ····················
G 1261 hCoV-19/USA/MN-MDH-1532/2020|EPI_ISL_527597|2020-0 ···················· ·A·················· ···················· ···················· ···················· ····················
G 1262 hCoV-19/USA/MN-MDH-1533/2020|EPI_ISL_527598|2020-0 ···················· ·A·················· ···················· ···················· ···················· ····················
G 1263 hCoV-19/USA/MN-MDH-1534/2020|EPI_ISL_527599|2020-0 ···················· ·A·················· ···················· ···················· ···················· ····················

| | | | | | | | |
|---|---|---|---|---|---|---|---|
| G | 1264 hCoV-19/England/BHRT-24B3521/2020\|EPI_ISL_528801\|2 | ···················· | ···················· | ····················· | ···················· | ···················· | ···················· |
| G | 1265 hCoV-19/England/BHRT-24B365B/2020\|EPI_ISL_528802\|2 | ···················· | ···················· | ····················· | ···················· | ···················· | ···················· |
| G | 1266 hCoV-19/England/BHRT-24B37EF/2020\|EPI_ISL_528803\|2 | ···················· | ···················· | ····················· | ···················· | ···················· | ···················· |
| G | 1267 hCoV-19/England/BHRT-24B35C7/2020\|EPI_ISL_528805\|2 | ···················· | ···················· | ····················· | ···················· | ···················· | ···················· |
| G | 1268 hCoV-19/England/BHRT-24B354F/2020\|EPI_ISL_528806\|2 | ···················· | ···················· | ····················· | ···················· | ···················· | ···················· |
| G | 1269 hCoV-19/England/BHRT-24B3794/2020\|EPI_ISL_528807\|2 | ···················· | ···················· | ····················· | ···················· | ···················· | ···················· |
| G | 1270 hCoV-19/England/BHRT-24B3819/2020\|EPI_ISL_528808\|2 | ···················· | ···················· | ····················· | ···················· | ···················· | ···················· |
| G | 1271 hCoV-19/India/TG-CCMB-X52/2020\|EPI_ISL_528860\|2020 | ···················· | ···················· | ····················· | ···················· | ···················· | ···················· |
| G | 1272 hCoV-19/India/TG-CCMB-X59/2020\|EPI_ISL_528861\|2020 | ···················· | ···················· | ····················· | ···················· | ···················· | ···················· |
| G | 1273 hCoV-19/India/TG-CCMB-X65/2020\|EPI_ISL_528862\|2020 | ···················· | ···················· | ····················· | ···················· | ···················· | ···················· |
| G | 1274 hCoV-19/India/TG-CCMB-X67/2020\|EPI_ISL_528863\|2020 | ···················· | ···················· | ····················· | ···················· | ···················· | ···················· |
| G | 1275 hCoV-19/India/TG-CCMB-X76/2020\|EPI_ISL_528864\|2020 | ···················· | ···················· | ····················· | ···················· | ···················· | ···················· |
| G | 1276 hCoV-19/India/TG-CCMB-X86/2020\|EPI_ISL_528865\|2020 | ···················· | ···················· | ····················· | ···················· | ···················· | ···················· |
| G | 1277 hCoV-19/India/TG-CCMB-X87/2020\|EPI_ISL_528866\|2020 | ···················· | ···················· | ····················· | ···················· | ···················· | ···················· |
| G | 1278 hCoV-19/India/TG-CCMB-X88/2020\|EPI_ISL_528867\|2020 | ···················· | ···················· | ····················· | ···················· | ···················· | ···················· |
| G | 1279 hCoV-19/India/TG-CCMB-X89/2020\|EPI_ISL_528868\|2020 | ···················· | ···················· | ····················· | ···················· | ···················· | ···················· |
| G | 1280 hCoV-19/India/TG-CCMB-X94/2020\|EPI_ISL_528869\|2020 | ···················· | ···················· | ····················· | ···················· | ···················· | ···················· |
| G | 1281 hCoV-19/England/LOND-125FBEE/2020\|EPI_ISL_516870\|2 | ···················· | ···················· | ····················· | ···················· | ···················· | ···················· |
| G | 1282 hCoV-19/England/LOND-125F814/2020\|EPI_ISL_516873\|2 | ···················· | ···················· | ····················· | ···················· | ···················· | ···················· |
| G | 1283 hCoV-19/England/LOND-125FAC3/2020\|EPI_ISL_516874\|2 | ···················· | ···················· | ····················· | ···················· | ···················· | ···················· |
| G | 1284 hCoV-19/England/LOND-125FD8E/2020\|EPI_ISL_516876\|2 | ···················· | ···················· | ····················· | ···················· | ···················· | ···················· |
| G | 1285 hCoV-19/England/LOND-125FD9D/2020\|EPI_ISL_516877\|2 | ···················· | ···················· | ····················· | ···················· | ···················· | ···················· |
| G | 1286 hCoV-19/England/LOND-125FE6D/2020\|EPI_ISL_516878\|2 | ···················· | ···················· | ····················· | ···················· | ···················· | ···················· |
| G | 1287 hCoV-19/Israel/CVL-n17051/2020\|EPI_ISL_516884\|2020 | ···················· | ···················· | ····················· | ···················· | ···················· | ···················· |
| G | 1288 hCoV-19/Israel/CVL-n17054/2020\|EPI_ISL_516885\|2020 | ···················· | ···················· | ····················· | ···················· | ···················· | ···················· |
| G | 1289 hCoV-19/Australia/VIC6408/2020\|EPI_ISL_521200\|2020 | ···················· | ···················· | ····················· | ···················· | ···················· | ···················· |
| G | 1290 hCoV-19/Australia/VIC6409/2020\|EPI_ISL_521201\|2020 | ···················· | ···················· | ····················· | ···················· | ···················· | ···················· |
| G | 1291 hCoV-19/Australia/VIC6410/2020\|EPI_ISL_521202\|2020 | ···················· | ···················· | ····················· | ···················· | ···················· | ···················· |
| G | 1292 hCoV-19/Australia/VIC6411/2020\|EPI_ISL_521203\|2020 | ···················· | ···················· | ····················· | ···················· | ···················· | ···················· |
| G | 1293 hCoV-19/Australia/VIC6412/2020\|EPI_ISL_521204\|2020 | ···················· | ···················· | ····················· | ···················· | ···················· | ···················· |
| G | 1294 hCoV-19/Australia/VIC6414/2020\|EPI_ISL_521206\|2020 | ···················· | ···················· | ····················· | ···················· | ···················· | ···················· |
| G | 1295 hCoV-19/Australia/VIC6415/2020\|EPI_ISL_521207\|2020 | ···················· | ···················· | ····················· | ···················· | ···················· | ···················· |
| G | 1296 hCoV-19/Australia/VIC6416/2020\|EPI_ISL_521208\|2020 | ···················· | ···················· | ····················· | ···················· | ···················· | ···················· |
| G | 1297 hCoV-19/Australia/VIC6419/2020\|EPI_ISL_521210\|2020 | ···················· | ···················· | ····················· | ···················· | ···················· | ···················· |
| G | 1298 hCoV-19/Australia/VIC6420/2020\|EPI_ISL_521211\|2020 | ···················· | ······A············· | ····················· | ···················· | ···················· | ···················· |
| G | 1299 hCoV-19/Australia/VIC6421/2020\|EPI_ISL_521212\|2020 | ···················· | ···················· | ····················· | ···················· | ···················· | ···················· |
| G | 1300 hCoV-19/Australia/VIC6422/2020\|EPI_ISL_521213\|2020 | ···················· | ···················· | ····················· | ···················· | ···················· | ···················· |
| G | 1301 hCoV-19/Australia/VIC6423/2020\|EPI_ISL_521214\|2020 | ···················· | ···················· | ····················· | ···················· | ···················· | ···················· |
| G | 1302 hCoV-19/Australia/VIC6424/2020\|EPI_ISL_521215\|2020 | ···················· | ···················· | ····················· | ···················· | ···················· | ···················· |
| G | 1303 hCoV-19/Australia/VIC6426/2020\|EPI_ISL_521217\|2020 | ···················· | ···················· | ····················· | ···················· | ···················· | ···················· |
| G | 1304 hCoV-19/Australia/VIC6427/2020\|EPI_ISL_521218\|2020 | ···················· | ···················· | ····················· | ···················· | ···················· | ···················· |
| G | 1305 hCoV-19/Australia/VIC6428/2020\|EPI_ISL_521219\|2020 | ···················· | ···················· | ····················· | ···················· | ···················· | ···················· |
| G | 1306 hCoV-19/Australia/VIC6429/2020\|EPI_ISL_521220\|2020 | ···················· | ···················· | ····················· | ···················· | ···················· | ···················· |
| G | 1307 hCoV-19/Australia/VIC6430/2020\|EPI_ISL_521221\|2020 | ···················· | ···················· | ····················· | ···················· | ···················· | ···················· |
| G | 1308 hCoV-19/Australia/VIC6431/2020\|EPI_ISL_521222\|2020 | ···················· | ···················· | ····················· | ···················· | ···················· | ···················· |
| G | 1309 hCoV-19/Australia/VIC6449/2020\|EPI_ISL_521227\|2020 | ···················· | ···················· | ····················· | ···················· | ···················· | ···················· |
| G | 1310 hCoV-19/Australia/VIC6450/2020\|EPI_ISL_521228\|2020 | ···················· | ···················· | ····················· | ···················· | ···················· | ···················· |
| G | 1311 hCoV-19/Australia/VIC6451/2020\|EPI_ISL_521229\|2020 | ···················· | ···················· | ····················· | ···················· | ···················· | ···················· |
| G | 1312 hCoV-19/Australia/VIC6452/2020\|EPI_ISL_521230\|2020 | ···················· | ···················· | ····················· | ···················· | ···················· | ···················· |
| G | 1313 hCoV-19/Australia/VIC6453/2020\|EPI_ISL_521231\|2020 | ···················· | ···················· | ····················· | ···················· | ···················· | ···················· |
| G | 1314 hCoV-19/Australia/VIC6456/2020\|EPI_ISL_521234\|2020 | ···················· | ······A············· | ····················· | ···················· | ···················· | ···················· |
| G | 1315 hCoV-19/Australia/VIC6457/2020\|EPI_ISL_521235\|2020 | ···················· | ······A············· | ····················· | ···················· | ···················· | ···················· |
| G | 1316 hCoV-19/Australia/VIC6458/2020\|EPI_ISL_521236\|2020 | ···················· | ···················· | ····················· | ···················· | ···················· | ···················· |
| G | 1317 hCoV-19/Australia/VIC6459/2020\|EPI_ISL_521237\|2020 | ···················· | ···················· | ····················· | ···················· | ···················· | ···················· |
| G | 1318 hCoV-19/Australia/VIC6460/2020\|EPI_ISL_521238\|2020 | ···················· | ···················· | ····················· | ···················· | ···················· | ···················· |
| G | 1319 hCoV-19/Australia/VIC6461/2020\|EPI_ISL_521239\|2020 | ···················· | ···················· | ····················· | ···················· | ···················· | ···················· |
| G | 1320 hCoV-19/Australia/VIC6462/2020\|EPI_ISL_521240\|2020 | ···················· | ······A············· | ····················· | ···················· | ···················· | ···················· |
| G | 1321 hCoV-19/Australia/VIC6463/2020\|EPI_ISL_521241\|2020 | ···················· | ······A············· | ····················· | ···················· | ···················· | ···················· |
| G | 1322 hCoV-19/Australia/VIC6464/2020\|EPI_ISL_521242\|2020 | ···················· | ···················· | ····················· | ···················· | ···················· | ···················· |
| G | 1323 hCoV-19/Australia/VIC6465/2020\|EPI_ISL_521243\|2020 | ···················· | ···················· | ····················· | ···················· | ···················· | ···················· |
| G | 1324 hCoV-19/Australia/VIC6466/2020\|EPI_ISL_521244\|2020 | ···················· | ···················· | ····················· | ···················· | ···················· | ···················· |
| G | 1325 hCoV-19/Australia/VIC6467/2020\|EPI_ISL_521245\|2020 | ···················· | ···················· | ····················· | ···················· | ···················· | ···················· |
| G | 1326 hCoV-19/Australia/VIC6468/2020\|EPI_ISL_521246\|2020 | ···················· | ···················· | ····················· | ···················· | ···················· | ···················· |
| G | 1327 hCoV-19/Australia/VIC6469/2020\|EPI_ISL_521247\|2020 | ···················· | ···················· | ····················· | ···················· | ···················· | ···················· |
| G | 1328 hCoV-19/Australia/VIC6470/2020\|EPI_ISL_521248\|2020 | ···················· | ···················· | ····················· | ···················· | ···················· | ···················· |
| G | 1329 hCoV-19/Australia/VIC6471/2020\|EPI_ISL_521249\|2020 | ···················· | ······A············· | ····················· | ···················· | ···················· | ···················· |
| G | 1330 hCoV-19/Australia/VIC6472/2020\|EPI_ISL_521250\|2020 | ···················· | ···················· | ····················· | ···················· | ···················· | ···················· |
| G | 1331 hCoV-19/Australia/VIC6473/2020\|EPI_ISL_521251\|2020 | ···················· | ···················· | ····················· | ···················· | ···················· | ···················· |
| G | 1332 hCoV-19/Australia/VIC6474/2020\|EPI_ISL_521252\|2020 | ···················· | ···················· | ····················· | ···················· | ···················· | ···················· |
| G | 1333 hCoV-19/Australia/VIC6475/2020\|EPI_ISL_521253\|2020 | ···················· | ···················· | ····················· | ···················· | ···················· | ···················· |
| G | 1334 hCoV-19/Australia/VIC6476/2020\|EPI_ISL_521254\|2020 | ···················· | ···················· | ····················· | ···················· | ···················· | ···················· |
| G | 1335 hCoV-19/Australia/VIC6477/2020\|EPI_ISL_521255\|2020 | ···················· | ···················· | ····················· | ···················· | ···················· | ···················· |
| G | 1336 hCoV-19/Australia/VIC6478/2020\|EPI_ISL_521256\|2020 | ···················· | ···················· | ····················· | ···················· | ···················· | ···················· |
| G | 1337 hCoV-19/Australia/VIC6479/2020\|EPI_ISL_521257\|2020 | ···················· | ···················· | ····················· | ···················· | ···················· | ···················· |
| G | 1338 hCoV-19/Australia/VIC6480/2020\|EPI_ISL_521258\|2020 | ···················· | ···················· | ····················· | ···················· | ···················· | ···················· |
| G | 1339 hCoV-19/Australia/VIC6482/2020\|EPI_ISL_521260\|2020 | ···················· | ···················· | ····················· | ···················· | ···················· | ···················· |
| G | 1340 hCoV-19/Australia/VIC6483/2020\|EPI_ISL_521261\|2020 | ···················· | ···················· | ····················· | ···················· | ···················· | ···················· |
| G | 1341 hCoV-19/Australia/VIC6484/2020\|EPI_ISL_521262\|2020 | ···················· | ···················· | ····················· | ···················· | ···················· | ···················· |
| G | 1342 hCoV-19/Australia/VIC6485/2020\|EPI_ISL_521263\|2020 | ···················· | ···················· | ····················· | ···················· | ···················· | ···················· |
| G | 1343 hCoV-19/Australia/VIC6486/2020\|EPI_ISL_521264\|2020 | ···················· | ···················· | ····················· | ···················· | ···················· | ···················· |

| | | | | | | | | |
|---|---|---|---|---|---|---|---|---|
| G | 1344 | hCoV-19/Australia/VIC6487/2020\|EPI_ISL_521265\|2020 | .................... | .................... | .................... | .................... | .................... | .................... |
| G | 1345 | hCoV-19/Australia/VIC6488/2020\|EPI_ISL_521266\|2020 | .................... | .................... | .................... | .................... | .................... | .................... |
| G | 1346 | hCoV-19/Australia/VIC6490/2020\|EPI_ISL_521268\|2020 | .................... | .................... | .................... | .................... | .................... | .................... |
| G | 1347 | hCoV-19/Australia/VIC6492/2020\|EPI_ISL_521269\|2020 | .................... | .................... | .................... | .................... | .................... | .................... |
| G | 1348 | hCoV-19/Australia/VIC6495/2020\|EPI_ISL_521270\|2020 | .................... | .................... | .................... | .................... | .................... | .................... |
| G | 1349 | hCoV-19/Australia/VIC6496/2020\|EPI_ISL_521271\|2020 | .................... | .................... | .................... | .................... | .................... | .................... |
| G | 1350 | hCoV-19/Australia/VIC6497/2020\|EPI_ISL_521272\|2020 | .................... | .................... | .................... | .................... | .................... | .................... |
| G | 1351 | hCoV-19/Australia/VIC6499/2020\|EPI_ISL_521274\|2020 | .................... | .................... | .................... | .................... | .................... | .................... |
| G | 1352 | hCoV-19/Australia/VIC6501/2020\|EPI_ISL_521275\|2020 | .................... | .................... | .................... | .................... | .................... | .................... |
| G | 1353 | hCoV-19/Australia/VIC6502/2020\|EPI_ISL_521276\|2020 | .................... | .................... | .................... | .................... | .................... | .................... |
| G | 1354 | hCoV-19/Australia/VIC6503/2020\|EPI_ISL_521277\|2020 | .................... | .................... | .................... | .................... | .................... | .................... |
| G | 1355 | hCoV-19/Australia/VIC6504/2020\|EPI_ISL_521278\|2020 | .................... | .................... | .................... | .................... | .................... | .................... |
| G | 1356 | hCoV-19/Australia/VIC6505/2020\|EPI_ISL_521279\|2020 | .................... | .................... | .................... | .................... | .................... | .................... |
| G | 1357 | hCoV-19/Australia/VIC6506/2020\|EPI_ISL_521280\|2020 | .................... | .................... | .................... | .................... | .................... | .................... |
| G | 1358 | hCoV-19/Australia/VIC6507/2020\|EPI_ISL_521281\|2020 | .................... | .................... | .................... | .................... | .................... | .................... |
| G | 1359 | hCoV-19/Australia/VIC6508/2020\|EPI_ISL_521282\|2020 | .................... | .................... | .................... | .................... | .................... | .................... |
| G | 1360 | hCoV-19/Australia/VIC6509/2020\|EPI_ISL_521283\|2020 | .................... | .................... | .................... | .................... | .................... | .................... |
| G | 1361 | hCoV-19/Australia/VIC6510/2020\|EPI_ISL_521284\|2020 | .................... | .................... | .................... | .................... | .................... | .................... |
| G | 1362 | hCoV-19/Australia/VIC6511/2020\|EPI_ISL_521285\|2020 | .................... | .................... | .................... | .................... | .................... | .................... |
| G | 1363 | hCoV-19/Australia/VIC6515/2020\|EPI_ISL_521287\|2020 | .................... | .................... | .................... | .................... | .................... | .................... |
| G | 1364 | hCoV-19/Australia/VIC6516/2020\|EPI_ISL_521288\|2020 | .................... | .................... | .................... | .................... | .................... | .................... |
| G | 1365 | hCoV-19/Australia/VIC6518/2020\|EPI_ISL_521289\|2020 | .................... | .................... | .................... | .................... | .................... | .................... |
| G | 1366 | hCoV-19/Australia/VIC6519/2020\|EPI_ISL_521290\|2020 | .................... | .................... | .................... | .................... | .................... | .................... |
| G | 1367 | hCoV-19/Australia/VIC6520/2020\|EPI_ISL_521291\|2020 | .................... | .................... | .................... | .................... | .................... | .................... |
| G | 1368 | hCoV-19/Australia/VIC6521/2020\|EPI_ISL_521292\|2020 | .................... | .................... | .................... | .................... | .................... | .................... |
| G | 1369 | hCoV-19/Australia/VIC6523/2020\|EPI_ISL_521293\|2020 | .................... | .................... | .................... | .................... | .................... | .................... |
| G | 1370 | hCoV-19/Australia/VIC6524/2020\|EPI_ISL_521294\|2020 | .................... | .................... | .................... | .................... | .................... | .................... |
| G | 1371 | hCoV-19/Australia/VIC6525/2020\|EPI_ISL_521295\|2020 | .................... | .................... | .................... | .................... | .................... | .................... |
| G | 1372 | hCoV-19/Australia/VIC6526/2020\|EPI_ISL_521296\|2020 | .................... | .................... | .................... | .................... | .................... | .................... |
| G | 1373 | hCoV-19/Australia/VIC6527/2020\|EPI_ISL_521297\|2020 | .................... | .................... | .................... | .................... | .................... | .................... |
| G | 1374 | hCoV-19/Australia/VIC6528/2020\|EPI_ISL_521298\|2020 | .................... | .................... | .................... | .................... | .................... | .................... |
| G | 1375 | hCoV-19/Australia/VIC6529/2020\|EPI_ISL_521299\|2020 | .................... | .................... | .................... | .................... | .................... | .................... |
| G | 1376 | hCoV-19/Australia/VIC6643/2020\|EPI_ISL_521400\|2020 | .................... | .................... | .................... | .................... | .................... | .................... |
| G | 1377 | hCoV-19/Australia/VIC6645/2020\|EPI_ISL_521402\|2020 | .................... | .................... | .................... | .................... | .................... | .................... |
| G | 1378 | hCoV-19/Australia/VIC6646/2020\|EPI_ISL_521403\|2020 | .................... | .................... | .................... | .................... | .................... | .................... |
| G | 1379 | hCoV-19/Australia/VIC6647/2020\|EPI_ISL_521404\|2020 | .................... | .................... | .................... | .................... | .................... | .................... |
| G | 1380 | hCoV-19/Australia/VIC6648/2020\|EPI_ISL_521405\|2020 | .................... | .................... | .................... | .................... | .................... | .................... |
| G | 1381 | hCoV-19/Australia/VIC6649/2020\|EPI_ISL_521406\|2020 | .................... | .................... | .................... | .................... | .................... | .................... |
| G | 1382 | hCoV-19/Australia/VIC6650/2020\|EPI_ISL_521407\|2020 | .................... | .................... | .................... | .................... | .................... | .................... |
| G | 1383 | hCoV-19/Australia/VIC6652/2020\|EPI_ISL_521409\|2020 | .................... | .................... | .................... | .................... | .................... | .................... |
| G | 1384 | hCoV-19/Australia/VIC6653/2020\|EPI_ISL_521410\|2020 | .................... | .................... | .................... | .................... | .................... | .................... |
| G | 1385 | hCoV-19/Australia/VIC6654/2020\|EPI_ISL_521411\|2020 | .................... | .................... | .................... | .................... | .................... | .................... |
| G | 1386 | hCoV-19/Australia/VIC6656/2020\|EPI_ISL_521412\|2020 | .................... | .................... | .................... | .................... | .................... | .................... |
| G | 1387 | hCoV-19/South Korea/KCDC2723/2020\|EPI_ISL_522502\|2 | .................... | .................... | .................... | .................... | .................... | .................... |
| G | 1388 | hCoV-19/South Korea/KCDC2724/2020\|EPI_ISL_522503\|2 | .................... | .................... | .................... | .................... | .................... | .................... |
| G | 1389 | hCoV-19/South Korea/KCDC2725/2020\|EPI_ISL_522504\|2 | .................... | .................... | .................... | .................... | .................... | .................... |
| G | 1390 | hCoV-19/South Korea/KCDC2726/2020\|EPI_ISL_522505\|2 | .................... | .................... | .................... | .................... | .................... | .................... |
| G | 1391 | hCoV-19/South Korea/KCDC2727/2020\|EPI_ISL_522506\|2 | .................... | .................... | .................... | .................... | .................... | .................... |
| G | 1392 | hCoV-19/South Korea/KCDC2728/2020\|EPI_ISL_522507\|2 | .................... | .................... | .................... | .................... | .................... | .................... |
| G | 1393 | hCoV-19/South Korea/KCDC2730/2020\|EPI_ISL_522509\|2 | .................... | .................... | .................... | .................... | .................... | .................... |
| G | 1394 | hCoV-19/South Korea/KCDC2731/2020\|EPI_ISL_522510\|2 | .................... | .................... | .................... | .................... | .................... | .................... |
| G | 1395 | hCoV-19/South Korea/KCDC2732/2020\|EPI_ISL_522511\|2 | .................... | .................... | .................... | .................... | .................... | .................... |
| G | 1396 | hCoV-19/South Korea/KCDC2733/2020\|EPI_ISL_522512\|2 | .................... | .................... | .................... | .................... | .................... | .................... |
| G | 1397 | hCoV-19/South Korea/KCDC2734/2020\|EPI_ISL_522513\|2 | .................... | .................... | .................... | .................... | .................... | .................... |
| G | 1398 | hCoV-19/South Korea/KCDC2735/2020\|EPI_ISL_522514\|2 | .................... | .................... | .................... | .................... | .................... | .................... |
| G | 1399 | hCoV-19/South Korea/KCDC2736/2020\|EPI_ISL_522515\|2 | .................... | .................... | .................... | .................... | .................... | .................... |
| G | 1400 | hCoV-19/South Korea/KCDC2737/2020\|EPI_ISL_522516\|2 | .................... | .................... | .................... | .................... | .................... | .................... |
| G | 1401 | hCoV-19/South Korea/KCDC2738/2020\|EPI_ISL_522517\|2 | .................... | .................... | .................... | .................... | .................... | .................... |
| G | 1402 | hCoV-19/South Korea/KCDC2739/2020\|EPI_ISL_522518\|2 | .................... | .................... | .................... | .................... | .................... | .................... |
| G | 1403 | hCoV-19/South Korea/KCDC2740/2020\|EPI_ISL_522519\|2 | .................... | .................... | .................... | .................... | .................... | .................... |
| G | 1404 | hCoV-19/South Korea/KCDC2741/2020\|EPI_ISL_522520\|2 | .................... | .................... | .................... | .................... | .................... | .................... |
| G | 1405 | hCoV-19/South Korea/KCDC2742/2020\|EPI_ISL_522521\|2 | .................... | .................... | .................... | .................... | .................... | .................... |
| G | 1406 | hCoV-19/South Korea/KCDC2743/2020\|EPI_ISL_522522\|2 | .................... | .................... | .................... | .................... | .................... | .................... |
| G | 1407 | hCoV-19/South Korea/KCDC2744/2020\|EPI_ISL_522523\|2 | .................... | .................... | .................... | .................... | .................... | .................... |
| G | 1408 | hCoV-19/South Korea/KCDC2745/2020\|EPI_ISL_522524\|2 | .................... | .................... | .................... | .................... | .................... | .................... |
| G | 1409 | hCoV-19/South Korea/KCDC2746/2020\|EPI_ISL_522525\|2 | .................... | .................... | .................... | .................... | .................... | .................... |
| G | 1410 | hCoV-19/South Korea/KCDC2747/2020\|EPI_ISL_522526\|2 | .................... | .................... | .................... | .................... | .................... | .................... |
| G | 1411 | hCoV-19/South Korea/KCDC2748/2020\|EPI_ISL_522527\|2 | .................... | .................... | .................... | .................... | .................... | .................... |
| G | 1412 | hCoV-19/South Korea/KCDC2749/2020\|EPI_ISL_522528\|2 | .................... | .................... | .................... | .................... | .................... | .................... |
| G | 1413 | hCoV-19/South Korea/KCDC2750/2020\|EPI_ISL_522529\|2 | .................... | .................... | .................... | .................... | .................... | .................... |
| G | 1414 | hCoV-19/South Korea/KCDC2751/2020\|EPI_ISL_522530\|2 | .................... | .................... | .................... | .................... | .................... | .................... |
| G | 1415 | hCoV-19/South Korea/KCDC2752/2020\|EPI_ISL_522531\|2 | .................... | .................... | .................... | .................... | .................... | .................... |
| G | 1416 | hCoV-19/South Korea/KCDC2753/2020\|EPI_ISL_522532\|2 | .................... | .................... | .................... | .................... | .................... | .................... |
| G | 1417 | hCoV-19/South Korea/KCDC2754/2020\|EPI_ISL_522533\|2 | .................... | .................... | .................... | .................... | .................... | .................... |
| G | 1418 | hCoV-19/South Korea/KCDC2755/2020\|EPI_ISL_522534\|2 | .................... | .................... | .................... | .................... | .................... | .................... |
| G | 1419 | hCoV-19/South Korea/KCDC2756/2020\|EPI_ISL_522535\|2 | .................... | .................... | .................... | .................... | .................... | .................... |
| G | 1420 | hCoV-19/South Korea/KCDC2757/2020\|EPI_ISL_522536\|2 | .................... | .................... | .................... | .................... | .................... | .................... |
| G | 1421 | hCoV-19/South Korea/KCDC2758/2020\|EPI_ISL_522537\|2 | .................... | .................... | .................... | .................... | .................... | .................... |
| G | 1422 | hCoV-19/South Korea/KCDC2759/2020\|EPI_ISL_522538\|2 | .................... | .................... | .................... | .................... | .................... | .................... |
| G | 1423 | hCoV-19/South Korea/KCDC2760/2020\|EPI_ISL_522539\|2 | .................... | .................... | .................... | .................... | .................... | .................... |

G 1424 hCoV-19/South Korea/KCDC2761/2020|EPI_ISL_522540|2 ···················· ···················· ···················· ···················· ···················· ····················
G 1425 hCoV-19/South Korea/KCDC2762/2020|EPI_ISL_522541|2 ···················· ···················· ···················· ···················· ···················· ····················
G 1426 hCoV-19/South Korea/KCDC2763/2020|EPI_ISL_522542|2 ···················· ···················· ···················· ···················· ···················· ····················
G 1427 hCoV-19/South Korea/KCDC2764/2020|EPI_ISL_522543|2 ···················· ···················· ···················· ···················· ···················· ····················
G 1428 hCoV-19/South Korea/KCDC2765/2020|EPI_ISL_522544|2 ···················· ···················· ···················· ···················· ···················· ····················
G 1429 hCoV-19/Reunion/RUN-PIMIT1/2020|EPI_ISL_522547|202 ···················· ···················· ···················· ···················· ···················· ····················
G 1430 hCoV-19/Reunion/RUN-PIMIT2/2020|EPI_ISL_522548|202 ···················· ···················· ···················· ···················· ···················· ····················
G 1431 hCoV-19/Reunion/RUN-PIMIT20/2020|EPI_ISL_522549|20 ···················· ···················· ···················· ···················· ···················· ····················
G 1432 hCoV-19/Reunion/RUN-PIMIT8/2020|EPI_ISL_522550|202 ···················· ···················· ···················· ···················· ···················· ····················
G 1444 hCoV-19/Australia/TAS271/2020|EPI_ISL_522700|2020- ···················· ···················· ···················· ···················· ···················· ····················
G 1445 hCoV-19/Australia/TAS272/2020|EPI_ISL_522701|2020- ···················· ···················· ···················· ···················· NNN················· ···················· 1
G 1446 hCoV-19/Australia/TAS273/2020|EPI_ISL_522702|2020- ···················· ···················· ···················· ···················· NNN················· ···············R···· 1
G 1448 hCoV-19/Australia/TAS278/2020|EPI_ISL_522705|2020- ···················· ···················· ···················· ···················· ···················· ····················
G 1457 hCoV-19/Switzerland/BL-ETHZ-240001/2020|EPI_ISL_52 ···················· ···················· ···················· ···················· ···················· ····················
G 1458 hCoV-19/Switzerland/GE-ETHZ-240002/2020|EPI_ISL_52 ···················· ···················· ···················· ···················· ···················· ····················
G 1459 hCoV-19/Switzerland/ZH-ETHZ-240003/2020|EPI_ISL_52 ···················· ···················· ···················· ···················· ···················· ····················
G 1460 hCoV-19/Switzerland/ZH-ETHZ-240004/2020|EPI_ISL_52 ···················· ···················· ···················· ···················· ···················· ····················
G 1461 hCoV-19/Switzerland/ZH-ETHZ-240005/2020|EPI_ISL_52 ···················· ···················· ···················· ···················· ···················· ····················
G 1462 hCoV-19/Switzerland/SG-ETHZ-240006/2020|EPI_ISL_52 ···················· ···················· ···················· ···················· ······K············· ····················
G 1463 hCoV-19/Switzerland/ZH-ETHZ-240007/2020|EPI_ISL_52 ···················· ···················· ···················· ···················· ···················· ····················
G 1464 hCoV-19/Switzerland/TI-ETHZ-240008/2020|EPI_ISL_52 ···················· ···················· ···················· ···················· ···················· ····················
G 1465 hCoV-19/Switzerland/AG-ETHZ-240009/2020|EPI_ISL_52 ···················· ···················· ···················· ···················· ···················· ····················
G 1466 hCoV-19/Switzerland/BL-ETHZ-240010/2020|EPI_ISL_52 ···················· ···················· ···················· ···················· ···················· ····················
G 1467 hCoV-19/Switzerland/BE-ETHZ-240015/2020|EPI_ISL_52 ···················· ·A·················· ···················· ···················· ···················· ····················
G 1468 hCoV-19/Switzerland/ZH-ETHZ-240016/2020|EPI_ISL_52 ···················· ···················· ···················· ···················· ···················· ····················
G 1469 hCoV-19/Switzerland/BL-ETHZ-240017/2020|EPI_ISL_52 ···················· ···················· ···················· ···················· ···················· ····················
G 1470 hCoV-19/Switzerland/BL-ETHZ-240018/2020|EPI_ISL_52 ···················· ···················· ···················· ···················· ···················· ····················
G 1471 hCoV-19/Switzerland/BL-ETHZ-240020/2020|EPI_ISL_52 ···················· ···················· ···················· ···················· ···················· ····················
G 1472 hCoV-19/Switzerland/BL-ETHZ-240021/2020|EPI_ISL_52 ···················· ···················· ···················· ···················· ···················· ····················
G 1473 hCoV-19/Switzerland/BL-ETHZ-240022/2020|EPI_ISL_52 ···················· ···················· ···················· ···················· ···················· ····················
G 1474 hCoV-19/Switzerland/BL-ETHZ-240023/2020|EPI_ISL_52 ···················· ···················· ···················· ···················· ···················· ····················
G 1475 hCoV-19/Switzerland/SG-ETHZ-240025/2020|EPI_ISL_52 ···················· ·A·················· ···················· ···················· ···················· ····················
G 1476 hCoV-19/Switzerland/SG-ETHZ-240028/2020|EPI_ISL_52 ···················· ·A·················· ···················· ···················· ···················· ····················
G 1477 hCoV-19/Switzerland/GE-ETHZ-240033/2020|EPI_ISL_52 ···················· ···················· ···················· ···················· ···················· ····················
G 1478 hCoV-19/Switzerland/ZH-ETHZ-240037/2020|EPI_ISL_52 ···················· ···················· ···················· ···················· ···················· ····················
G 1479 hCoV-19/Switzerland/ZH-ETHZ-240038/2020|EPI_ISL_52 ···················· ···················· ···················· ···················· ···················· ····················
G 1480 hCoV-19/Switzerland/ZH-ETHZ-240039/2020|EPI_ISL_52 ···················· ···················· ···················· ···················· ···················· ····················
G 1481 hCoV-19/Switzerland/BL-ETHZ-240041/2020|EPI_ISL_52 ···················· ···················· ···················· ···················· ···················· ····················
G 1482 hCoV-19/Switzerland/ZH-ETHZ-240042/2020|EPI_ISL_52 ···················· ···················· ···················· ···················· ······K············· ····················
G 1483 hCoV-19/Switzerland/GE-ETHZ-240043/2020|EPI_ISL_52 ···················· ···················· ···················· ···················· ···················· ····················
G 1484 hCoV-19/Switzerland/BL-ETHZ-240045/2020|EPI_ISL_52 ···················· ···················· ···················· ···················· ···················· ····················
G 1485 hCoV-19/Switzerland/BL-ETHZ-240046/2020|EPI_ISL_52 ···················· ···················· ···················· ···················· ···················· ····················
G 1486 hCoV-19/Switzerland/ZH-ETHZ-240047/2020|EPI_ISL_52 ···················· ···················· ···················· ···················· ···················· ····················
G 1487 hCoV-19/Switzerland/BL-ETHZ-240050/2020|EPI_ISL_52 ···················· ···················· ···················· ···················· ···················· ····················
G 1488 hCoV-19/Switzerland/ZH-ETHZ-240051/2020|EPI_ISL_52 ···················· ···················· ···················· ···················· ···················· ····················
G 1489 hCoV-19/Switzerland/ZH-ETHZ-240053/2020|EPI_ISL_52 ···················· ···················· ···················· ···················· ···················· ····················
G 1490 hCoV-19/Switzerland/ZH-ETHZ-240054/2020|EPI_ISL_52 ···················· ···················· ···················· ···················· ···················· ····················
G 1491 hCoV-19/Switzerland/BE-ETHZ-240055/2020|EPI_ISL_52 ···················· ···················· ···················· ···················· ···················· ····················
G 1492 hCoV-19/Switzerland/GE-ETHZ-240058/2020|EPI_ISL_52 ···················· ···················· ···················· ···················· ···················· ····················
G 1493 hCoV-19/Switzerland/GE-ETHZ-240059/2020|EPI_ISL_52 ···················· ···················· ···················· ···················· ···················· ····················
G 1494 hCoV-19/Canada/ON-UHTC_0139/2020|EPI_ISL_526275|20 ···················· ···················· ···················· ···················· ···················· ····················
G 1495 hCoV-19/Canada/ON-UHTC_0151/2020|EPI_ISL_526284|20 ···················· ···················· ···················· ···················· ···················· ····················
G 1496 hCoV-19/England/ALDP-97B526/2020|EPI_ISL_526288|20 ···················· ···················· ···················· ···················· ···················· ····················
G 1497 hCoV-19/England/ALDP-97B580/2020|EPI_ISL_526289|20 ···················· ···················· ···················· ···················· ···················· ····················
G 1498 hCoV-19/England/ALDP-97B623/2020|EPI_ISL_526290|20 ···················· ···················· ···················· ···················· ···················· ····················
G 1499 hCoV-19/England/ALDP-97B896/2020|EPI_ISL_526291|20 ···················· ···················· ···················· ···················· ···················· ····················
G 1500 hCoV-19/England/ALDP-97BDF1/2020|EPI_ISL_526294|20 ···················· ···················· ···················· ···················· ···················· ····················
G 1501 hCoV-19/England/ALDP-97BFA0/2020|EPI_ISL_526295|20 ···················· ···················· ···················· ···················· ···················· ····················
G 1502 hCoV-19/England/ALDP-97BFBF/2020|EPI_ISL_526296|20 ···················· ···················· ···················· ···················· ···················· ····················
G 1503 hCoV-19/England/ALDP-97BFFB/2020|EPI_ISL_526298|20 ···················· ···················· ···················· ···················· ···················· ····················
G 1504 hCoV-19/United Arab Emirates/434/2020|EPI_ISL_5287 ···················· ···················· ···················· ···················· ···················· ····················
G 1505 hCoV-19/United Arab Emirates/44/2020|EPI_ISL_52870 ···················· ···················· ···················· ···················· ···················· ····················
G 1506 hCoV-19/United Arab Emirates/484/2020|EPI_ISL_5287 ···················· ···················· ···················· ···················· ···················· ····················
G 1507 hCoV-19/United Arab Emirates/529/2020|EPI_ISL_5287 ···················· ···················· ···················· ···················· ···················· ····················
G 1508 hCoV-19/United Arab Emirates/54/2020|EPI_ISL_52870 ···················· ···················· ···················· ···················· ···················· ····················
G 1509 hCoV-19/United Arab Emirates/558/2020|EPI_ISL_5287 ···················· ·A·················· ···················· ···················· ···················· ····················
G 1510 hCoV-19/United Arab Emirates/56/2020|EPI_ISL_52870 ···················· ···················· ···················· ···················· ···················· ····················
G 1511 hCoV-19/United Arab Emirates/581/2020|EPI_ISL_5287 ···················· ·A·················· ···················· ···················· ···················· ····················
G 1512 hCoV-19/United Arab Emirates/C2-109/2020|EPI_ISL_5 ···················· ···················· ···················· ···················· ···················· ····················
G 1513 hCoV-19/United Arab Emirates/H1/2020|EPI_ISL_52870 ···················· ···················· ···················· ···················· ···················· ····················
G 1514 hCoV-19/United Arab Emirates/H10/2020|EPI_ISL_5287 ···················· ···················· ···················· ···················· ···················· ····················
G 1515 hCoV-19/United Arab Emirates/H12/2020|EPI_ISL_5287 ···················· ···················· ···················· ···················· ···················· ····················
G 1516 hCoV-19/United Arab Emirates/H13/2020|EPI_ISL_5287 ···················· ···················· ···················· ···················· ···················· ····················
G 1517 hCoV-19/United Arab Emirates/H18/2020|EPI_ISL_5287 ···················· ···················· ···················· ···················· ···················· ····················
G 1518 hCoV-19/United Arab Emirates/H2/2020|EPI_ISL_52871 ···················· ···················· ···················· ···················· ···················· ····················
G 1519 hCoV-19/United Arab Emirates/H20/2020|EPI_ISL_5287 ···················· ···················· ···················· ···················· ···················· ····················
G 1520 hCoV-19/United Arab Emirates/H21/2020|EPI_ISL_5287 ···················· ···················· ···················· ···················· ···················· ····················
G 1521 hCoV-19/United Arab Emirates/H23/2020|EPI_ISL_5287 ···················· ···················· ···················· ···················· ···················· ····················
G 1522 hCoV-19/United Arab Emirates/H27/2020|EPI_ISL_5287 ···················· ···················· ···················· ···················· ···················· ····················
G 1523 hCoV-19/United Arab Emirates/H5/2020|EPI_ISL_52871 ···················· ···················· ···················· ···················· ···················· ····················

| | | | | | | | |
|---|---|---|---|---|---|---|---|
| G | 1524 hCoV-19/United Arab Emirates/H8/2020\|EPI_ISL_52872 | .................... | .................... | .................... | .................... | .................... | .................... |
| G | 1525 hCoV-19/United Arab Emirates/H9/2020\|EPI_ISL_52872 | .................... | .................... | .................... | .................... | .................... | .................... |
| G | 1526 hCoV-19/India/TG-CCMB-X115/2020\|EPI_ISL_528849\|202 | .................... | .................... | .................... | .................... | .................... | .................... |
| G | 1527 hCoV-19/India/TG-CCMB-X116/2020\|EPI_ISL_528850\|202 | .................... | .................... | .................... | .................... | .................... | .................... |
| G | 1528 hCoV-19/India/TG-CCMB-X14/2020\|EPI_ISL_528851\|2020 | .................... | .................... | .................... | .................... | .................... | .................... |
| G | 1529 hCoV-19/India/TG-CCMB-X15/2020\|EPI_ISL_528852\|2020 | .................... | .................... | .................... | .................... | .................... | .................... |
| G | 1530 hCoV-19/India/TG-CCMB-X16/2020\|EPI_ISL_528853\|2020 | .................... | .................... | .................... | .................... | .................... | .................... |
| G | 1531 hCoV-19/India/TG-CCMB-X42/2020\|EPI_ISL_528854\|2020 | .................... | .................... | .................... | .................... | .................... | .................... |
| G | 1532 hCoV-19/India/TG-CCMB-X43/2020\|EPI_ISL_528855\|2020 | .................... | .................... | .................... | .................... | .................... | .................... |
| G | 1533 hCoV-19/India/TG-CCMB-X44/2020\|EPI_ISL_528856\|2020 | .................... | .................... | .................... | .................... | .................... | .................... |
| G | 1534 hCoV-19/India/TG-CCMB-X46/2020\|EPI_ISL_528857\|2020 | .................... | .................... | .................... | .................... | .................... | .................... |
| G | 1535 hCoV-19/India/TG-CCMB-X49/2020\|EPI_ISL_528858\|2020 | .................... | .................... | .................... | .................... | .................... | .................... |
| G | 1536 hCoV-19/India/TG-CCMB-X50/2020\|EPI_ISL_528859\|2020 | .................... | .................... | .................... | .................... | .................... | .................... |
| G | 1537 hCoV-19/South Korea/KCDC2653/2020\|EPI_ISL_516783\|2 | .................... | .................... | .................... | .................... | .................... | .................... |
| G | 1538 hCoV-19/South Korea/KCDC2654/2020\|EPI_ISL_516784\|2 | .................... | .................... | .................... | .................... | .................... | .................... |
| G | 1539 hCoV-19/South Korea/KCDC2655/2020\|EPI_ISL_516785\|2 | .................... | .................... | .................... | .................... | .................... | .................... |
| G | 1540 hCoV-19/Hong Kong/HKU-200823-001/2020\|EPI_ISL_5167 | .................... | .................... | .................... | .................... | .................... | .................... |
| G | 1541 hCoV-19/Hong Kong/HKU-200823-002/2020\|EPI_ISL_5167 | .................... | .................... | .................... | .................... | .................... | .................... |
| G | 1542 hCoV-19/Australia/VIC6481/2020\|EPI_ISL_521259\|2020 | .................... | .................... | .................... | .................... | .................... | .................... |
| G | 1543 hCoV-19/Australia/VIC6489/2020\|EPI_ISL_521267\|2020 | .................... | .................... | .................... | .................... | .................... | .................... |
| G | 1544 hCoV-19/Australia/TAS15/2020\|EPI_ISL_522610\|2020-0 | .................... | .................... | .................... | .................... | .................... | .................... |
| G | 1545 hCoV-19/Switzerland/ZH-ETHZ-240060/2020\|EPI_ISL_52 | .................... | .................... | .................... | .................... | .................... | .................... |
| G | 1546 hCoV-19/Switzerland/BE-ETHZ-240061/2020\|EPI_ISL_52 | .................... | .A.................. | .................... | .................... | .................... | .................... |
| G | 1547 hCoV-19/Switzerland/BL-ETHZ-240062/2020\|EPI_ISL_52 | .................... | .................... | .................... | .................... | .................... | .................... |
| G | 1548 hCoV-19/Switzerland/ZH-ETHZ-240063/2020\|EPI_ISL_52 | .................... | .................... | .................... | .................... | .................... | .................... |
| G | 1549 hCoV-19/Switzerland/BE-ETHZ-240065/2020\|EPI_ISL_52 | .................... | .................... | .................... | .................... | .................... | .................... |
| G | 1550 hCoV-19/Switzerland/BE-ETHZ-240066/2020\|EPI_ISL_52 | .................... | .................... | .................... | .................... | .................... | .................... |
| G | 1551 hCoV-19/Switzerland/BS-ETHZ-240069/2020\|EPI_ISL_52 | .................... | .................... | .................... | .................... | .................... | .................... |
| G | 1552 hCoV-19/Switzerland/ZH-ETHZ-240070/2020\|EPI_ISL_52 | .................... | .A.................. | .................... | .................... | .................... | .................... |
| G | 1553 hCoV-19/Switzerland/ZH-ETHZ-240071/2020\|EPI_ISL_52 | .................... | .................... | .................... | .................... | .................... | .................... |
| G | 1554 hCoV-19/Switzerland/BL-ETHZ-240072/2020\|EPI_ISL_52 | .................... | .................... | .................... | .................... | .................... | .................... |
| G | 1555 hCoV-19/Switzerland/SO-ETHZ-240073/2020\|EPI_ISL_52 | .................... | .A.................. | .................... | .................... | .................... | .................... |
| G | 1556 hCoV-19/Switzerland/ZH-ETHZ-240075/2020\|EPI_ISL_52 | .................... | .................... | .................... | .................... | .................... | .................... |
| G | 1557 hCoV-19/Switzerland/UR-ETHZ-240077/2020\|EPI_ISL_52 | .................... | .A.................. | .................... | .................... | .................... | .................... |
| G | 1558 hCoV-19/Switzerland/UR-ETHZ-240078/2020\|EPI_ISL_52 | .................... | .A.................. | .................... | .................... | .................... | .................... |
| G | 1559 hCoV-19/Switzerland/GE-ETHZ-240079/2020\|EPI_ISL_52 | .................... | .................... | .................... | .................... | .................... | .................... |
| G | 1560 hCoV-19/Switzerland/BE-ETHZ-240080/2020\|EPI_ISL_52 | .................... | .................... | .................... | .................... | .................... | .................... |
| G | 1561 hCoV-19/Switzerland/UR-ETHZ-240082/2020\|EPI_ISL_52 | .................... | .A.................. | .................... | .................... | .................... | .................... |
| G | 1562 hCoV-19/Switzerland/VS-ETHZ-240083/2020\|EPI_ISL_52 | .................... | .................... | .................... | .................... | .................... | .................... |
| G | 1563 hCoV-19/Switzerland/BL-ETHZ-240084/2020\|EPI_ISL_52 | .................... | .................... | .................... | .................... | .................... | .................... |
| G | 1564 hCoV-19/Switzerland/BE-ETHZ-240086/2020\|EPI_ISL_52 | .................... | .A.................. | .................... | .................... | .................... | .................... |
| G | 1565 hCoV-19/Switzerland/UR-ETHZ-240087/2020\|EPI_ISL_52 | .................... | .................... | .................... | .................... | .................... | .................... |
| G | 1566 hCoV-19/Switzerland/BE-ETHZ-240089/2020\|EPI_ISL_52 | .................... | .................... | .................... | .................... | .................... | .................... |
| G | 1567 hCoV-19/Switzerland/BS-ETHZ-240090/2020\|EPI_ISL_52 | .................... | .................... | .................... | .................... | .................... | .................... |
| G | 1568 hCoV-19/Switzerland/ZH-ETHZ-240091/2020\|EPI_ISL_52 | .................... | .A.................. | .................... | .................... | .................... | .................... |
| G | 1569 hCoV-19/Germany/NW-HHU-56/2020\|EPI_ISL_523927\|2020 | .................... | .................... | .................... | .................... | .................... | .................... |
| G | 1570 hCoV-19/Germany/NW-HHU-58/2020\|EPI_ISL_523929\|2020 | .................... | .................... | .................... | .................... | .................... | .................... |
| G | 1571 hCoV-19/Germany/NW-HHU-60/2020\|EPI_ISL_523931\|2020 | .................... | .A.................. | .................... | .................... | .................... | .................... |
| G | 1572 hCoV-19/Germany/NW-HHU-61/2020\|EPI_ISL_523932\|2020 | .................... | .................... | .................... | .................... | .................... | .................... |
| G | 1573 hCoV-19/Germany/NW-HHU-62/2020\|EPI_ISL_523933\|2020 | .................... | .................... | .................... | .................... | .................... | .................... |
| G | 1574 hCoV-19/Germany/NW-HHU-63/2020\|EPI_ISL_523934\|2020 | .................... | .................... | .................... | .................... | .................... | .................... |
| G | 1575 hCoV-19/Germany/NW-HHU-64/2020\|EPI_ISL_523935\|2020 | .................... | .................... | .................... | .................... | .................... | .................... |
| G | 1576 hCoV-19/Germany/NW-HHU-66/2020\|EPI_ISL_523937\|2020 | .................... | .................... | .................... | .................... | .................... | .................... |
| G | 1577 hCoV-19/Germany/NW-HHU-67/2020\|EPI_ISL_523938\|2020 | .................... | .................... | .................... | .................... | .................... | .................... |
| G | 1578 hCoV-19/Germany/NW-HHU-68/2020\|EPI_ISL_523939\|2020 | .................... | .................... | .................... | .................... | .................... | .................... |
| G | 1579 hCoV-19/Germany/NW-HHU-69/2020\|EPI_ISL_523940\|2020 | .................... | .................... | .................... | .................... | .................... | .................... |
| G | 1580 hCoV-19/Germany/NW-HHU-70/2020\|EPI_ISL_523941\|2020 | .................... | .................... | .................... | .................... | .................... | .................... |
| G | 1581 hCoV-19/Germany/NW-HHU-71/2020\|EPI_ISL_523942\|2020 | .................... | .................... | .................... | .................... | .................... | .................... |
| G | 1582 hCoV-19/Germany/NW-HHU-72/2020\|EPI_ISL_523943\|2020 | .................... | .................... | .................... | .................... | .................... | .................... |
| G | 1583 hCoV-19/Germany/NW-HHU-73/2020\|EPI_ISL_523944\|2020 | .................... | .................... | .................... | .................... | .................... | .................... |
| G | 1584 hCoV-19/Germany/NW-HHU-74/2020\|EPI_ISL_523945\|2020 | .................... | .................... | .................... | .................... | .................... | .................... |
| G | 1585 hCoV-19/Germany/NW-HHU-76/2020\|EPI_ISL_523947\|2020 | .................... | .................... | .................... | .................... | .................... | .................... |
| G | 1586 hCoV-19/Germany/NW-HHU-77/2020\|EPI_ISL_523948\|2020 | .................... | .................... | .................... | .................... | .................... | .................... |
| G | 1587 hCoV-19/Germany/NW-HHU-78/2020\|EPI_ISL_523949\|2020 | .................... | .................... | .................... | .................... | .................... | .................... |
| G | 1588 hCoV-19/Germany/NW-HHU-79/2020\|EPI_ISL_523950\|2020 | .................... | .................... | .................... | .................... | .................... | .................... |
| G | 1589 hCoV-19/England/203380362/2020\|EPI_ISL_523951\|2020 | .................... | .................... | .................... | .................... | .................... | .................... |
| G | 1590 hCoV-19/United Arab Emirates/L3290260632/2020\|EPI_ | .................... | .................... | .................... | .................... | .................... | .................... |
| G | 1591 hCoV-19/USA/WA-OHSU-0957/2020\|EPI_ISL_526100\|2020- | .................... | .................... | .................... | .................... | .................... | .................... |
| G | 1592 hCoV-19/USA/WA-OHSU-0973/2020\|EPI_ISL_526101\|2020- | .................... | .................... | .................... | .................... | .................... | .................... |
| G | 1593 hCoV-19/USA/WA-OHSU-0987/2020\|EPI_ISL_526102\|2020- | .................... | .................... | .................... | .................... | .................... | .................... |
| G | 1594 hCoV-19/USA/WA-OHSU-1571/2020\|EPI_ISL_526103\|2020- | .................... | .................... | .................... | .................... | .................... | .................... |
| G | 1595 hCoV-19/USA/WA-OHSU-1611/2020\|EPI_ISL_526104\|2020- | .................... | .A.................. | .................... | .................... | .................... | .................... |
| G | 1596 hCoV-19/USA/WA-OHSU-1647/2020\|EPI_ISL_526105\|2020- | .................... | .................... | .................... | .................... | .................... | .................... |
| G | 1597 hCoV-19/USA/WA-OHSU-1661/2020\|EPI_ISL_526106\|2020- | .................... | .................... | .................... | .................... | .................... | .................... |
| G | 1598 hCoV-19/USA/WA-OHSU-1721/2020\|EPI_ISL_526107\|2020- | .................... | .A.................. | .................... | .................... | .................... | .................... |
| G | 1599 hCoV-19/USA/WA-OHSU-1799/2020\|EPI_ISL_526108\|2020- | .................... | .................... | .................... | .................... | .................... | .................... |
| G | 1600 hCoV-19/USA/WA-OHSU-1831/2020\|EPI_ISL_526109\|2020- | .................... | .A.................. | .................... | .................... | .................... | .................... |
| G | 1601 hCoV-19/USA/WA-OHSU-1909/2020\|EPI_ISL_526110\|2020- | .................... | .A.................. | .................... | .................... | .................... | .................... |
| G | 1602 hCoV-19/USA/WA-OHSU-1933/2020\|EPI_ISL_526111\|2020- | .................... | .................... | .................... | .................... | .................... | .................... |
| G | 1603 hCoV-19/USA/WA-OHSU-1953/2020\|EPI_ISL_526112\|2020- | .................... | .A.................. | .................... | .................... | .................... | .................... |

G 1604 hCoV-19/USA/WA-OHSU-1965/2020|EPI_ISL_526113|2020- .................... .................... .................... .................... .................... ....................
G 1605 hCoV-19/USA/WA-OHSU-1967/2020|EPI_ISL_526114|2020- .................... .................... .................... .................... .................... ....................
G 1606 hCoV-19/Australia/NSW773/2020|EPI_ISL_526115|2020- .................... .................... .................... .................... .................... ....................
G 1607 hCoV-19/Australia/NSW774/2020|EPI_ISL_526116|2020- .................... .................... .................... .................... .................... ....................
G 1608 hCoV-19/Australia/NSW775/2020|EPI_ISL_526117|2020- .................... .................... .................... .................... .................... ....................
G 1609 hCoV-19/Australia/NSW776/2020|EPI_ISL_526118|2020- .................... .................... .................... .................... .................... ....................
G 1610 hCoV-19/Australia/NSW777/2020|EPI_ISL_526119|2020- .................... .................... .................... .................... .................... ....................
G 1611 hCoV-19/Australia/NSW778/2020|EPI_ISL_526120|2020- .................... .................... .................... .................... .................... ....................
G 1612 hCoV-19/Australia/NSW779/2020|EPI_ISL_526121|2020- .................... .................... .................... .................... .................... ....................
G 1613 hCoV-19/Australia/NSW780/2020|EPI_ISL_526122|2020- .................... .................... .................... .................... .................... ....................
G 1614 hCoV-19/Australia/NSW781/2020|EPI_ISL_526123|2020- .................... .................... .................... .................... .................... ....................
G 1615 hCoV-19/Australia/NSW782/2020|EPI_ISL_526124|2020- .................... .................... .................... .................... .................... ....................
G 1616 hCoV-19/Australia/NSW783/2020|EPI_ISL_526125|2020- .................... .................... .................... .................... .................... ....................
G 1617 hCoV-19/Australia/NSW784/2020|EPI_ISL_526126|2020- .................... .................... .................... .................... .................... ....................
G 1618 hCoV-19/Australia/NSW785/2020|EPI_ISL_526127|2020- .................... .................... .................... .................... .................... ....................
G 1619 hCoV-19/Australia/NSW786/2020|EPI_ISL_526128|2020- .................... .................... .................... .................... .................... ....................
G 1620 hCoV-19/Australia/NSW787/2020|EPI_ISL_526129|2020- .................... .................... .................... .................... .................... ....................
G 1621 hCoV-19/Australia/NSW788/2020|EPI_ISL_526130|2020- .................... .................... .................... .................... .................... ....................
G 1622 hCoV-19/Australia/NSW789/2020|EPI_ISL_526131|2020- .................... .................... .................... .................... .................... ....................
G 1623 hCoV-19/Australia/NSW790/2020|EPI_ISL_526132|2020- .................... .................... .................... .................... .................... ....................
G 1624 hCoV-19/Australia/NSW791/2020|EPI_ISL_526133|2020- .................... .................... .................... .................... .................... ....................
G 1625 hCoV-19/Australia/NSW792/2020|EPI_ISL_526134|2020- .................... .................... .................... .................... .................... ....................
G 1626 hCoV-19/Australia/NSW793/2020|EPI_ISL_526135|2020- .................... .................... .................... .................... .................... ....................
G 1627 hCoV-19/Australia/NSW794/2020|EPI_ISL_526136|2020- .................... .................... .................... .................... .................... ....................
G 1628 hCoV-19/Australia/NSW795/2020|EPI_ISL_526137|2020- .................... .................... .................... .................... .................... ....................
G 1629 hCoV-19/Australia/NSW796/2020|EPI_ISL_526138|2020- .................... .................... .................... .................... .................... ....................
G 1630 hCoV-19/Australia/NSW797/2020|EPI_ISL_526139|2020- .................... .................... .................... .................... .................... ....................
G 1631 hCoV-19/Australia/NSW798/2020|EPI_ISL_526140|2020- .................... .................... .................... .................... .................... ....................
G 1632 hCoV-19/Australia/NSW799/2020|EPI_ISL_526141|2020- .................... .................... .................... .................... .................... ....................
G 1633 hCoV-19/Australia/NSW800/2020|EPI_ISL_526142|2020- .................... .................... .................... .................... .................... ....................
G 1634 hCoV-19/Australia/NSW801/2020|EPI_ISL_526143|2020- .................... .................... .................... .................... .................... ....................
G 1635 hCoV-19/Australia/NSW802/2020|EPI_ISL_526144|2020- .................... .................... .................... .................... .................... ....................
G 1636 hCoV-19/Australia/NSW803/2020|EPI_ISL_526145|2020- .................... .................... .................... .................... .................... ....................
G 1637 hCoV-19/Australia/NSW804/2020|EPI_ISL_526146|2020- .................... .................... .................... .................... .................... ....................
G 1638 hCoV-19/Australia/NSW805/2020|EPI_ISL_526147|2020- .................... .................... .................... .................... .................... ....................
G 1639 hCoV-19/Australia/NSW806/2020|EPI_ISL_526148|2020- .................... .................... .................... .................... .................... ....................
G 1640 hCoV-19/Australia/NSW807/2020|EPI_ISL_526149|2020- .................... .................... .................... .................... .................... ....................
G 1641 hCoV-19/Australia/NSW808/2020|EPI_ISL_526150|2020- .................... .................... .................... .................... .................... ....................
G 1642 hCoV-19/Australia/NSW811/2020|EPI_ISL_526153|2020- .................... .................... .................... .................... .................... ....................
G 1643 hCoV-19/Australia/NSW812/2020|EPI_ISL_526154|2020- .................... .................... .................... .................... .................... ....................
G 1644 hCoV-19/Australia/NSW813/2020|EPI_ISL_526155|2020- .................... .................... .................... .................... .................... ....................
G 1645 hCoV-19/Australia/NSW814/2020|EPI_ISL_526156|2020- .................... .................... .................... .................... .................... ....................
G 1646 hCoV-19/Australia/NSW818/2020|EPI_ISL_526160|2020- .................... .................... .................... .................... .................... ....................
G 1647 hCoV-19/Australia/NSW819/2020|EPI_ISL_526161|2020- .................... .................... .................... .................... .................... ....................
G 1648 hCoV-19/Australia/NSW820/2020|EPI_ISL_526162|2020- .................... .................... .................... .................... .................... ....................
G 1649 hCoV-19/Australia/NSW821/2020|EPI_ISL_526163|2020- .................... .................... .................... .................... .................... ....................
G 1650 hCoV-19/Australia/NSW822/2020|EPI_ISL_526164|2020- .................... .................... .................... .................... .................... ....................
G 1651 hCoV-19/Australia/NSW823/2020|EPI_ISL_526165|2020- .................... .................... .................... .................... .................... ....................
G 1652 hCoV-19/Australia/NSW824/2020|EPI_ISL_526166|2020- .................... .................... .................... .................... .................... ....................
G 1653 hCoV-19/Australia/NSW825/2020|EPI_ISL_526167|2020- .................... .................... .................... .................... .................... ....................
G 1654 hCoV-19/Australia/NSW826/2020|EPI_ISL_526168|2020- .................... .................... .................... .................... .................... ....................
G 1655 hCoV-19/Australia/NSW827/2020|EPI_ISL_526169|2020- .................... .................... .................... .................... .................... ....................
G 1656 hCoV-19/Australia/NSW828/2020|EPI_ISL_526170|2020- .................... .................... .................... .................... .................... ....................
G 1657 hCoV-19/Australia/NSW829/2020|EPI_ISL_526171|2020- .................... .................... .................... .................... .................... ....................
G 1658 hCoV-19/Australia/NSW830/2020|EPI_ISL_526172|2020- .................... .................... .................... .................... .................... ....................
G 1659 hCoV-19/Australia/NSW831/2020|EPI_ISL_526173|2020- .................... .................... .................... .................... .................... ....................
G 1660 hCoV-19/Australia/NSW832/2020|EPI_ISL_526174|2020- .................... .................... .................... .................... .................... ....................
G 1661 hCoV-19/Australia/NSW833/2020|EPI_ISL_526175|2020- .................... .................... .................... .................... .................... ....................
G 1662 hCoV-19/Australia/NSW834/2020|EPI_ISL_526176|2020- .................... .................... .................... .................... .................... ....................
G 1663 hCoV-19/Australia/NSW835/2020|EPI_ISL_526177|2020- .................... .................... .................... .................... .................... ....................
G 1664 hCoV-19/Australia/NSW836/2020|EPI_ISL_526178|2020- .................... .................... .................... .................... .................... ....................
G 1665 hCoV-19/Australia/NSW837/2020|EPI_ISL_526179|2020- .................... .................... .................... .................... .................... ....................
G 1666 hCoV-19/Australia/NSW838/2020|EPI_ISL_526180|2020- .................... .................... .................... .................... .................... ....................
G 1667 hCoV-19/Australia/NSW839/2020|EPI_ISL_526181|2020- .................... .................... .................... .................... .................... ....................
G 1668 hCoV-19/Australia/NSW840/2020|EPI_ISL_526182|2020- .................... .................... .................... .................... .................... ....................
G 1669 hCoV-19/Australia/NSW841/2020|EPI_ISL_526183|2020- .................... .................... .................... .................... .................... ....................
G 1670 hCoV-19/Australia/NSW842/2020|EPI_ISL_526184|2020- .................... .................... .................... .................... .................... ....................
G 1671 hCoV-19/Australia/NSW843/2020|EPI_ISL_526185|2020- .................... .................... .................... .................... .................... ....................
G 1672 hCoV-19/Australia/NSW844/2020|EPI_ISL_526186|2020- .................... .................... .................... .................... .................... ....................
G 1673 hCoV-19/Australia/NSW845/2020|EPI_ISL_526187|2020- .................... .................... .................... .................... .................... ....................
G 1674 hCoV-19/Australia/NSW846/2020|EPI_ISL_526188|2020- .................... .................... .................... .................... .................... ....................
G 1675 hCoV-19/Australia/NSW847/2020|EPI_ISL_526189|2020- .................... .................... .................... .................... .................... ....................
G 1676 hCoV-19/Australia/NSW848/2020|EPI_ISL_526190|2020- .................... .................... .................... .................... .................... ....................
G 1677 hCoV-19/Australia/NSW849/2020|EPI_ISL_526191|2020- .................... .................... .................... .................... .................... ....................
G 1678 hCoV-19/Australia/NSW850/2020|EPI_ISL_526192|2020- .................... .................... .................... .................... .................... ....................
G 1679 hCoV-19/Australia/NSW851/2020|EPI_ISL_526193|2020- .................... .................... .................... .................... .................... ....................
G 1680 hCoV-19/Australia/NSW852/2020|EPI_ISL_526194|2020- .................... .................... .................... .................... .................... ....................
G 1681 hCoV-19/Australia/NSW853/2020|EPI_ISL_526195|2020- .................... .................... .................... .................... .................... ....................
G 1682 hCoV-19/Australia/NSW854/2020|EPI_ISL_526196|2020- .................... .................... .................... .................... .................... ....................
G 1683 hCoV-19/Australia/NSW855/2020|EPI_ISL_526197|2020- .................... .................... .................... .................... .................... ....................

| | | | | | | | |
|---|---|---|---|---|---|---|---|
| G | 1684 hCoV-19/Australia/NSW856/2020\|EPI_ISL_526198\|2020- | .................... | .................... | ..................... | .................... | .................... | .................... |
| G | 1685 hCoV-19/Australia/NSW857/2020\|EPI_ISL_526199\|2020- | .................... | .................... | ..................... | .................... | .................... | .................... |
| G | 1686 hCoV-19/Australia/NSW2718/2020\|EPI_ISL_527400\|2020 | .................... | .................... | ..................... | .................... | .................... | .................... |
| G | 1687 hCoV-19/USA/CO-CSU-B1397/2020\|EPI_ISL_527401\|2020- | .................... | .................... | ..................... | .................... | .................... | .................... |
| G | 1688 hCoV-19/USA/CO-CSU-C0363-1/2020\|EPI_ISL_527402\|202 | .................... | .................... | ..................... | .................... | .................... | .................... |
| G | 1689 hCoV-19/USA/CO-CSU-C0363-2/2020\|EPI_ISL_527403\|202 | .................... | .................... | ..................... | .................... | .................... | .................... |
| G | 1690 hCoV-19/USA/CO-CSU-C0768/2020\|EPI_ISL_527404\|2020- | .................... | .................... | ..................... | .................... | .................... | .................... |
| G | 1691 hCoV-19/USA/CO-CSU-C0881-1/2020\|EPI_ISL_527405\|202 | .................... | .................... | ..................... | .................... | .................... | .................... |
| G | 1692 hCoV-19/USA/CO-CSU-C0881-2/2020\|EPI_ISL_527406\|202 | .................... | .................... | ..................... | .................... | .................... | .................... |
| G | 1693 hCoV-19/USA/CO-CSU-C1255/2020\|EPI_ISL_527407\|2020- | .................... | .................... | ..................... | .................... | .................... | .................... |
| G | 1694 hCoV-19/USA/CO-CSU-C2673-1/2020\|EPI_ISL_527408\|202 | .................... | .................... | ..................... | .................... | .................... | .................... |
| G | 1695 hCoV-19/USA/CO-CSU-C2673-2/2020\|EPI_ISL_527409\|202 | .................... | .................... | ..................... | .................... | .................... | .................... |
| G | 1696 hCoV-19/USA/CO-CSU-C2980-1/2020\|EPI_ISL_527410\|202 | .................... | ......A............. | ..................... | .................... | .................... | .................... |
| G | 1697 hCoV-19/USA/CO-CSU-C2980-2/2020\|EPI_ISL_527411\|202 | .................... | ......A............. | ..................... | .................... | .................... | .................... |
| G | 1698 hCoV-19/USA/CO-CSU-D0204/2020\|EPI_ISL_527412\|2020- | .................... | .................... | ..................... | .................... | .................... | .................... |
| G | 1699 hCoV-19/USA/CO-CSU-D0259/2020\|EPI_ISL_527413\|2020- | .................... | .................... | ..................... | .................... | .................... | .................... |
| G | 1700 hCoV-19/USA/CO-CSU-D0290/2020\|EPI_ISL_527414\|2020- | .................... | .................... | ..................... | .................... | .................... | .................... |
| G | 1701 hCoV-19/USA/CO-CSU-D0376/2020\|EPI_ISL_527415\|2020- | .................... | .................... | ..................... | .................... | .................... | .................... |
| G | 1702 hCoV-19/USA/CO-CSU-D0459-1/2020\|EPI_ISL_527416\|202 | .................... | .................... | ..................... | .................... | .................... | .................... |
| G | 1703 hCoV-19/USA/CO-CSU-D0459-2/2020\|EPI_ISL_527417\|202 | .................... | .................... | ..................... | .................... | .................... | .................... |
| G | 1704 hCoV-19/USA/CO-CSU-D0459-3/2020\|EPI_ISL_527418\|202 | .................... | .................... | ..................... | .................... | .................... | .................... |
| G | 1705 hCoV-19/USA/CO-CSU-D0590-1/2020\|EPI_ISL_527419\|202 | .................... | .................... | ..................... | .................... | .................... | .................... |
| G | 1706 hCoV-19/USA/CO-CSU-D0590-2/2020\|EPI_ISL_527420\|202 | .................... | .................... | ..................... | .................... | .................... | .................... |
| G | 1707 hCoV-19/USA/CO-CSU-D0769-1/2020\|EPI_ISL_527421\|202 | .................... | .................... | ..................... | .................... | .................... | .................... |
| G | 1708 hCoV-19/USA/CO-CSU-D0769-2/2020\|EPI_ISL_527422\|202 | .................... | .................... | ..................... | .................... | .................... | .................... |
| G | 1709 hCoV-19/USA/CO-CSU-D0769-3/2020\|EPI_ISL_527423\|202 | .................... | .................... | ..................... | .................... | .................... | .................... |
| G | 1710 hCoV-19/USA/CO-CSU-D0789-1/2020\|EPI_ISL_527424\|202 | .................... | .................... | ..................... | .................... | .................... | .................... |
| G | 1711 hCoV-19/USA/CO-CSU-D0789-2/2020\|EPI_ISL_527425\|202 | .................... | .................... | ..................... | .................... | .................... | .................... |
| G | 1712 hCoV-19/USA/CO-CSU-D1050-1/2020\|EPI_ISL_527426\|202 | .................... | .................... | ..................... | .................... | .................... | .................... |
| G | 1713 hCoV-19/USA/CO-CSU-D1050-2/2020\|EPI_ISL_527427\|202 | .................... | .................... | ..................... | .................... | .................... | .................... |
| G | 1714 hCoV-19/USA/CO-CSU-D1057-1/2020\|EPI_ISL_527428\|202 | .................... | .................... | ..................... | .................... | .................... | .................... |
| G | 1715 hCoV-19/USA/CO-CSU-D1057-2/2020\|EPI_ISL_527429\|202 | .................... | .................... | ..................... | .................... | .................... | .................... |
| G | 1716 hCoV-19/USA/CO-CSU-D1057-3/2020\|EPI_ISL_527430\|202 | .................... | .................... | ..................... | .................... | .................... | .................... |
| G | 1717 hCoV-19/USA/CO-CSU-D1060-1/2020\|EPI_ISL_527431\|202 | .................... | .................... | ..................... | .................... | .................... | .................... |
| G | 1718 hCoV-19/USA/CO-CSU-D1060-2/2020\|EPI_ISL_527432\|202 | .................... | .................... | ....K................ | .................... | .................... | .................... |
| G | 1719 hCoV-19/USA/CO-CSU-D1068/2020\|EPI_ISL_527433\|2020- | .................... | .................... | ..................... | .................... | .................... | .................... |
| G | 1720 hCoV-19/USA/CO-CSU-D1075-1/2020\|EPI_ISL_527434\|202 | .................... | .................... | ..................... | .................... | .................... | .................... |
| G | 1721 hCoV-19/USA/CO-CSU-D1075-2/2020\|EPI_ISL_527435\|202 | .................... | .................... | ..................... | .................... | .................... | .................... |
| G | 1722 hCoV-19/USA/CO-CSU-D1085-1/2020\|EPI_ISL_527436\|202 | .................... | .................... | ..................... | .................... | .................... | .................... |
| G | 1723 hCoV-19/USA/CO-CSU-D1085-2/2020\|EPI_ISL_527437\|202 | .................... | .................... | ..................... | .................... | .................... | .................... |
| G | 1724 hCoV-19/USA/CO-CSU-D1085-3/2020\|EPI_ISL_527438\|202 | .................... | .................... | ..................... | .................... | .................... | .................... |
| G | 1725 hCoV-19/USA/CO-CSU-D1490-1/2020\|EPI_ISL_527439\|202 | .................... | .................... | ..................... | .................... | .................... | .................... |
| G | 1726 hCoV-19/USA/CO-CSU-D1490-2/2020\|EPI_ISL_527440\|202 | .................... | .................... | ..................... | .................... | .................... | .................... |
| G | 1727 hCoV-19/USA/CO-CSU-D1493-1/2020\|EPI_ISL_527441\|202 | .................... | ......A............. | ..................... | .................... | .................... | .................... |
| G | 1728 hCoV-19/USA/CO-CSU-D1657-1/2020\|EPI_ISL_527442\|202 | .................... | .................... | ..................... | .................... | .................... | .................... |
| G | 1729 hCoV-19/USA/CO-CSU-D1657-2/2020\|EPI_ISL_527443\|202 | .................... | .................... | ..................... | .................... | .................... | .................... |
| G | 1730 hCoV-19/USA/CO-CSU-D1657-3/2020\|EPI_ISL_527444\|202 | .................... | .................... | ..................... | .................... | .................... | .................... |
| G | 1731 hCoV-19/USA/CO-CSU-D1665/2020\|EPI_ISL_527445\|2020- | .................... | .................... | ..................... | .................... | .................... | .................... |
| G | 1732 hCoV-19/USA/CO-CSU-D1675/2020\|EPI_ISL_527446\|2020- | .................... | .................... | ..................... | .................... | .................... | .................... |
| G | 1733 hCoV-19/USA/CO-CSU-D1694/2020\|EPI_ISL_527447\|2020- | .................... | .................... | ..................... | .................... | .................... | .................... |
| G | 1734 hCoV-19/USA/CO-CSU-D1696/2020\|EPI_ISL_527448\|2020- | .................... | .................... | ..................... | .................... | .................... | .................... |
| G | 1735 hCoV-19/USA/CO-CSU-D1773/2020\|EPI_ISL_527449\|2020- | .................... | .................... | ..................... | .................... | .................... | .................... |
| G | 1736 hCoV-19/USA/CO-CSU-D1862/2020\|EPI_ISL_527450\|2020- | .................... | .................... | ..................... | .................... | .................... | .................... |
| G | 1737 hCoV-19/USA/CO-CSU-D1882-1/2020\|EPI_ISL_527451\|202 | .................... | .................... | ..................... | .................... | .................... | .................... |
| G | 1738 hCoV-19/USA/CO-CSU-D1882-2/2020\|EPI_ISL_527452\|202 | .................... | .................... | ..................... | .................... | .................... | .................... |
| G | 1739 hCoV-19/USA/CO-CSU-D1977/2020\|EPI_ISL_527453\|2020- | .................... | .................... | ..................... | .................... | .................... | .................... |
| G | 1740 hCoV-19/USA/CO-CSU-D1990-1/2020\|EPI_ISL_527454\|202 | .................... | .................... | ..................... | .................... | .................... | .................... |
| G | 1741 hCoV-19/USA/CO-CSU-D1990-2/2020\|EPI_ISL_527455\|202 | .................... | .................... | ..................... | .................... | .................... | .................... |
| G | 1742 hCoV-19/USA/CO-CSU-D2094/2020\|EPI_ISL_527456\|2020- | .................... | .................... | ..................... | .................... | .................... | .................... |
| G | 1743 hCoV-19/USA/CO-CSU-D2099/2020\|EPI_ISL_527457\|2020- | .................... | .................... | ..................... | .................... | .................... | .................... |
| G | 1744 hCoV-19/USA/CO-CSU-D2259/2020\|EPI_ISL_527458\|2020- | .................... | .................... | ..................... | .................... | .................... | .................... |
| G | 1745 hCoV-19/USA/CO-CSU-D2282-1/2020\|EPI_ISL_527459\|202 | .................... | .................... | ..................... | .................... | .................... | .................... |
| G | 1746 hCoV-19/USA/CO-CSU-D2282-2/2020\|EPI_ISL_527460\|202 | .................... | .................... | ..................... | .................... | .................... | .................... |
| G | 1747 hCoV-19/USA/CO-CSU-D2282-3/2020\|EPI_ISL_527462\|202 | .................... | .................... | ..................... | .................... | .................... | .................... |
| G | 1748 hCoV-19/USA/CO-CSU-D2373-1/2020\|EPI_ISL_527463\|202 | .................... | .................... | ..................... | .................... | .................... | .................... |
| G | 1749 hCoV-19/USA/CO-CSU-D2373-2/2020\|EPI_ISL_527464\|202 | .................... | .................... | ..................... | .................... | .................... | .................... |
| G | 1750 hCoV-19/USA/CO-CSU-D2399-1/2020\|EPI_ISL_527465\|202 | .................... | .................... | ..................... | .................... | .................... | .................... |
| G | 1751 hCoV-19/USA/CO-CSU-D2399-2/2020\|EPI_ISL_527466\|202 | .................... | .................... | ..................... | .................... | .................... | .................... |
| G | 1752 hCoV-19/USA/CO-CSU-D2591-1/2020\|EPI_ISL_527468\|202 | .................... | .................... | ..................... | .................... | .................... | .................... |
| G | 1753 hCoV-19/USA/CO-CSU-D2591-2/2020\|EPI_ISL_527469\|202 | .................... | .................... | ..................... | .................... | .................... | .................... |
| G | 1754 hCoV-19/USA/CO-CSU-D2598-1/2020\|EPI_ISL_527470\|202 | .................... | .................... | ..................... | .................... | .................... | .................... |
| G | 1755 hCoV-19/USA/CO-CSU-D2598-2/2020\|EPI_ISL_527471\|202 | .................... | .................... | ..................... | .................... | .................... | .................... |
| G | 1756 hCoV-19/USA/CO-CSU-D2674/2020\|EPI_ISL_527472\|2020- | .................... | ......A............. | ..................... | .................... | .................... | .................... |
| G | 1757 hCoV-19/USA/CO-CSU-D2678/2020\|EPI_ISL_527473\|2020- | .................... | .................... | ..................... | .................... | .................... | .................... |
| G | 1758 hCoV-19/USA/CO-CSU-D2680/2020\|EPI_ISL_527474\|2020- | .................... | ......A............. | ..................... | .................... | .................... | .................... |
| G | 1759 hCoV-19/USA/CO-CSU-D3084-1/2020\|EPI_ISL_527475\|202 | .................... | .................... | ..................... | .................... | .................... | .................... |
| G | 1760 hCoV-19/USA/CO-CSU-D3084-2/2020\|EPI_ISL_527476\|202 | .................... | .................... | ..................... | .................... | .................... | .................... |
| G | 1761 hCoV-19/USA/CO-CSU-D3171/2020\|EPI_ISL_527477\|2020- | .................... | .................... | ..................... | .................... | .................... | .................... |
| G | 1762 hCoV-19/USA/CO-CSU-D3376/2020\|EPI_ISL_527478\|2020- | .................... | .................... | ..................... | .................... | .................... | .................... |
| G | 1763 hCoV-19/USA/CO-CSU-E0462-1/2020\|EPI_ISL_527479\|202 | .................... | .................... | ..................... | .................... | .................... | .................... |

| | | | | | | | | |
|---|---|---|---|---|---|---|---|---|
| G | 1764 hCoV-19/USA/CO-CSU-E0462-2/2020\|EPI_ISL_527480\|202 | ···················· | ···················· | ···················· | ···················· | ···················· | ···················· | |
| G | 1765 hCoV-19/USA/CO-CSU-E0468/2020\|EPI_ISL_527481\|2020- | ···················· | ···················· | ···················· | ···················· | ···················· | ···················· | |
| G | 1766 hCoV-19/USA/CO-CSU-E0983-2/2020\|EPI_ISL_527484\|202 | ···················· | ···················· | ···················· | ···················· | ···················· | ···················· | |
| G | 1767 hCoV-19/USA/CO-CSU-E0983-3/2020\|EPI_ISL_527485\|202 | ···················· | ···················· | ···················· | ···················· | ···················· | ···················· | |
| G | 1768 hCoV-19/USA/CO-CSU-E1302/2020\|EPI_ISL_527486\|2020- | ···················· | ···················· | ···················· | ···················· | ···················· | ···················· | |
| G | 1769 hCoV-19/USA/CO-CSU-E2267/2020\|EPI_ISL_527487\|2020- | ···················· | ···················· | ···················· | ···················· | ···················· | ···················· | |
| G | 1770 hCoV-19/Malaysia/MGI-G873/2020\|EPI_ISL_528738\|2020 | ···················· | ···················· | ···················· | ···················· | ···················· | ···················· | |
| G | 1771 hCoV-19/Malaysia/MGI-M32/2020\|EPI_ISL_528739\|2020- | ···················· | ···················· | ···················· | ···················· | ···················· | ···················· | |
| G | 1772 hCoV-19/Malaysia/MGI-M64/2020\|EPI_ISL_528740\|2020- | ···················· | ···················· | ···················· | ···················· | ···················· | ···················· | |
| G | 1773 hCoV-19/Malaysia/MGI-M71/2020\|EPI_ISL_528741\|2020- | ···················· | ···················· | ···················· | ···················· | ···················· | ···················· | |
| G | 1774 hCoV-19/Malaysia/MGI-M76/2020\|EPI_ISL_528742\|2020- | ···················· | ···················· | ···················· | ···················· | ···················· | ···················· | |
| G | 1775 hCoV-19/Malaysia/MGI-MAEPS54/2020\|EPI_ISL_528743\|2 | ···················· | ···················· | ···················· | ···················· | ···················· | ···················· | |
| G | 1776 hCoV-19/Malaysia/MGI-MAEPS67/2020\|EPI_ISL_528744\|2 | ···················· | ···················· | ···················· | ···················· | ···················· | ···················· | |
| G | 1777 hCoV-19/Indonesia/JB-TFRIC19-R24754/2020\|EPI_ISL_5 | ···················· | ···················· | ···················· | ···················· | ···················· | ···················· | |
| G | 1778 hCoV-19/Indonesia/JB-TFRIC19-R47609/2020\|EPI_ISL_5 | ···················· | ···················· | ···················· | ···················· | ···················· | ···················· | |
| G | 1779 hCoV-19/Indonesia/JB-TFRIC19-R30535/2020\|EPI_ISL_5 | ···················· | ···················· | ···················· | ···················· | ···················· | ···················· | |
| G | 1780 hCoV-19/Indonesia/JB-TFRIC19-R48562/2020\|EPI_ISL_5 | ···················· | ···················· | ···················· | ···················· | ···················· | ···················· | |
| G | 1781 hCoV-19/Indonesia/JB-TFRIC19-R4975/2020\|EPI_ISL_52 | ···················· | ···················· | ···················· | ···················· | ···················· | ···················· | |
| G | 1782 hCoV-19/Indonesia/JB-TFRIC19-R49542/2020\|EPI_ISL_5 | ···················· | ···················· | ···················· | ···················· | ···················· | ···················· | |
| G | 1783 hCoV-19/Indonesia/JB-TFRIC19-R49344/2020\|EPI_ISL_5 | ···················· | ···················· | ···················· | ···················· | ···················· | ···················· | |
| G | 1784 hCoV-19/Indonesia/JB-TFRIC19-R48875/2020\|EPI_ISL_5 | ···················· | ···A················ | ···················· | ···················· | ···················· | ···················· | |
| G | 1785 hCoV-19/Indonesia/JB-TFRIC19-R53817/2020\|EPI_ISL_5 | ···················· | ···················· | ···················· | ···················· | ···················· | ···················· | |
| G | 1786 hCoV-19/Indonesia/JB-TFRIC19-R49544/2020\|EPI_ISL_5 | ···················· | ···················· | ···················· | ···················· | ···················· | ···················· | |
| G | 1787 hCoV-19/England/BHRT-24B35A9/2020\|EPI_ISL_528788\|2 | ···················· | ···················· | ···················· | ···················· | ···················· | ···················· | |
| G | 1788 hCoV-19/England/BHRT-24B3925/2020\|EPI_ISL_528790\|2 | ···················· | ···················· | ···················· | ···················· | ···················· | ···················· | |
| G | 1789 hCoV-19/England/BHRT-24B3A13/2020\|EPI_ISL_528791\|2 | ···················· | ···················· | ···················· | ···················· | ···················· | ···················· | |
| G | 1790 hCoV-19/England/BHRT-24B358B/2020\|EPI_ISL_528793\|2 | ···················· | ···················· | ···················· | ···················· | ···················· | ···················· | |
| G | 1791 hCoV-19/England/BHRT-24B36E2/2020\|EPI_ISL_528796\|2 | ···················· | ···················· | ···················· | ···················· | ···················· | ···················· | |
| G | 1792 hCoV-19/England/BHRT-24B3679/2020\|EPI_ISL_528799\|2 | ···················· | ···················· | ···················· | ···················· | ···················· | ···················· | |
| G | 1793 hCoV-19/USA/FL-BPHL-1014/2020\|EPI_ISL_517904\|2020- | ···················· | ···················· | ···················· | ···················· | ···················· | ···················· | |
| G | 1794 hCoV-19/USA/FL-BPHL-1015/2020\|EPI_ISL_517905\|2020- | ···················· | ···················· | ···················· | ···················· | ···················· | ···················· | |
| G | 1795 hCoV-19/USA/FL-BPHL-1016/2020\|EPI_ISL_517906\|2020- | ···················· | ···················· | ···················· | ···················· | ···················· | ···················· | |
| G | 1796 hCoV-19/USA/FL-BPHL-1017/2020\|EPI_ISL_517907\|2020- | ···················· | ···················· | ···················· | ···················· | ···················· | ···················· | |
| G | 1797 hCoV-19/USA/FL-BPHL-1018/2020\|EPI_ISL_517908\|2020- | ···················· | ···················· | ···················· | ···················· | ···················· | ···················· | |
| G | 1798 hCoV-19/USA/FL-BPHL-1019/2020\|EPI_ISL_517909\|2020- | ···················· | ···················· | ···················· | ···················· | ···················· | ···················· | |
| G | 1799 hCoV-19/USA/FL-BPHL-1022/2020\|EPI_ISL_517912\|2020- | ···················· | ·············A······ | ···················· | ···················· | ···················· | ···················· | |
| G | 1801 hCoV-19/USA/FL-BPHL-1025/2020\|EPI_ISL_517915\|2020- | ···················· | ···················· | ···················· | ···················· | ···················· | ···················· | |
| G | 1802 hCoV-19/USA/FL-BPHL-1026/2020\|EPI_ISL_517916\|2020- | ···················· | ···················· | ···················· | ···················· | ···················· | ···················· | |
| G | 1803 hCoV-19/USA/FL-BPHL-1028/2020\|EPI_ISL_517918\|2020- | ···················· | ···················· | ···················· | ···················· | ···················· | ···················· | |
| G | 1804 hCoV-19/USA/FL-BPHL-1030/2020\|EPI_ISL_517920\|2020- | ···················· | ···················· | ···················· | ···················· | ···················· | ···················· | |
| G | 1805 hCoV-19/USA/FL-BPHL-1031/2020\|EPI_ISL_517921\|2020- | ···················· | ···················· | ···················· | ···················· | ···················· | ···················· | |
| G | 1806 hCoV-19/USA/FL-BPHL-1032/2020\|EPI_ISL_517922\|2020- | ···················· | ···················· | ···················· | ···················· | ···················· | ···················· | |
| G | 1807 hCoV-19/USA/FL-BPHL-1033/2020\|EPI_ISL_517923\|2020- | ···················· | ···················· | ···················· | ···················· | ···················· | ···················· | |
| G | 1808 hCoV-19/USA/FL-BPHL-1034/2020\|EPI_ISL_517924\|2020- | ···················· | ···················· | ···················· | ···················· | ···················· | ···················· | |
| G | 1809 hCoV-19/USA/FL-BPHL-1035/2020\|EPI_ISL_517925\|2020- | ···················· | ···················· | ···················· | ···················· | ···················· | ···················· | |
| G | 1810 hCoV-19/USA/FL-BPHL-1036/2020\|EPI_ISL_517926\|2020- | ···················· | ···················· | ···················· | ···················· | ···················· | ···················· | |
| G | 1811 hCoV-19/USA/FL-BPHL-1038/2020\|EPI_ISL_517928\|2020- | ···················· | ···················· | ···················· | ···················· | ···················· | ···················· | |
| G | 1812 hCoV-19/USA/FL-BPHL-1039/2020\|EPI_ISL_517929\|2020- | ···················· | ···················· | ···················· | ···················· | ···················· | ···················· | |
| G | 1814 hCoV-19/USA/FL-BPHL-1041/2020\|EPI_ISL_517931\|2020- | ···················· | ···················· | ···················· | ···················· | ···················· | ···················· | |
| G | 1815 hCoV-19/USA/FL-BPHL-1042/2020\|EPI_ISL_517932\|2020- | ···················· | ···················· | ···················· | ···················· | ···················· | ···················· | |
| G | 1816 hCoV-19/USA/FL-BPHL-1043/2020\|EPI_ISL_517933\|2020- | ···················· | ···················· | ···················· | ···················· | ···················· | ···················· | |
| G | 1817 hCoV-19/USA/FL-BPHL-1044/2020\|EPI_ISL_517934\|2020- | ···················· | ···················· | ···················· | ···················· | ···················· | ···················· | |
| G | 1820 hCoV-19/USA/FL-BPHL-1049/2020\|EPI_ISL_517939\|2020- | ···················· | ···················· | ···················· | ···················· | ···················· | ···················· | |
| G | 1821 hCoV-19/USA/FL-BPHL-1050/2020\|EPI_ISL_517940\|2020- | ···················· | ···················· | ···················· | ···················· | ···················· | ···················· | |
| G | 1822 hCoV-19/USA/FL-BPHL-1051/2020\|EPI_ISL_517941\|2020- | ···················· | ···················· | ···················· | ···················· | ···················· | ···················· | |
| G | 1823 hCoV-19/USA/FL-BPHL-1054/2020\|EPI_ISL_517944\|2020- | ···················· | ···················· | ···················· | ···················· | ···················· | ···············A···· | |
| G | 1824 hCoV-19/USA/FL-BPHL-1055/2020\|EPI_ISL_517945\|2020- | ···················· | ···T················ | ···················· | ···················· | ···················· | ···················· | |
| G | 1825 hCoV-19/USA/FL-BPHL-1056/2020\|EPI_ISL_517946\|2020- | ···················· | ···················· | ···················· | ···················· | ···················· | ···················· | |
| G | 1827 hCoV-19/USA/FL-BPHL-1059/2020\|EPI_ISL_517949\|2020- | ···················· | ···················· | ···················· | ···················· | ···················· | ···················· | |
| G | 1828 hCoV-19/USA/FL-BPHL-1060/2020\|EPI_ISL_517950\|2020- | ···················· | ···················· | ···················· | ···················· | ···················· | ···················· | |
| G | 1829 hCoV-19/USA/FL-BPHL-1061/2020\|EPI_ISL_517951\|2020- | ···················· | ···················· | ···················· | ···················· | ···················· | ···················· | |
| G | 1830 hCoV-19/USA/FL-BPHL-1062/2020\|EPI_ISL_517952\|2020- | ···················· | ···················· | ···················· | ···················· | ···················· | ···················· | |
| G | 1831 hCoV-19/USA/FL-BPHL-1063/2020\|EPI_ISL_517953\|2020- | ···················· | ···················· | ···················· | ···················· | ···················· | ···················· | |
| G | 1832 hCoV-19/USA/FL-BPHL-1064/2020\|EPI_ISL_517954\|2020- | ···················· | ···················· | ···················· | ···················· | ···················· | ···················· | |
| G | 1833 hCoV-19/USA/FL-BPHL-1066/2020\|EPI_ISL_517956\|2020- | ···················· | ···················· | ···················· | ···················· | ···················· | ···················· | |
| G | 1834 hCoV-19/Peru/CUS-INS-068/2020\|EPI_ISL_517958\|2020- | ···················· | ···················· | ···················· | ···················· | ···················· | ···················· | |
| G | 1837 hCoV-19/Australia/VIC3433/2020\|EPI_ISL_521102\|2020 | ···················· | ···················· | ···················· | ···················· | ···················· | ···················· | |
| G | 1838 hCoV-19/Australia/VIC3435/2020\|EPI_ISL_521104\|2020 | ···················· | ···················· | ···················· | ···················· | ···················· | ···················· | |
| G | 1839 hCoV-19/Australia/VIC3438/2020\|EPI_ISL_521105\|2020 | ···················· | ···················· | ···················· | ···················· | ···················· | ···················· | |
| G | 1841 hCoV-19/Australia/VIC3441/2020\|EPI_ISL_521107\|2020 | ···················· | ···················· | ···················· | ···················· | ···················· | ···················· | |
| G | 1842 hCoV-19/Australia/VIC3442/2020\|EPI_ISL_521108\|2020 | ···················· | ···················· | ···················· | ···················· | ···················· | ···················· | |
| G | 1843 hCoV-19/Australia/VIC3444/2020\|EPI_ISL_521110\|2020 | ···················· | ···················· | ···················· | ···················· | ···················· | ···················· | |
| G | 1845 hCoV-19/Australia/VIC3446/2020\|EPI_ISL_521112\|2020 | ···················· | ···················· | ···················· | ···················· | ···················· | ···················· | |
| G | 1846 hCoV-19/Australia/VIC3448/2020\|EPI_ISL_521114\|2020 | ···················· | ···················· | ···················· | ···················· | ···················· | ···················· | |
| G | 1847 hCoV-19/Australia/VIC3449/2020\|EPI_ISL_521115\|2020 | ···················· | ···················· | ···················· | ···················· | ···················· | ···················· | |
| G | 1848 hCoV-19/Australia/VIC3451/2020\|EPI_ISL_521116\|2020 | ···················· | ···················· | ···················· | ···················· | ···················· | ···················· | |
| G | 1849 hCoV-19/Australia/VIC3452/2020\|EPI_ISL_521117\|2020 | ···················· | ···················· | ···················· | ···················· | ···················· | ···················· | |
| G | 1850 hCoV-19/Australia/VIC3454/2020\|EPI_ISL_521118\|2020 | ···················· | ···················· | ···················· | ···················· | ···················· | ···················· | |
| G | 1851 hCoV-19/Australia/VIC3455/2020\|EPI_ISL_521119\|2020 | ···················· | ···················· | ···················· | ···················· | NNN················· | ···················· | 1 |
| G | 1852 hCoV-19/Australia/VIC3456/2020\|EPI_ISL_521120\|2020 | ···················· | ···················· | ···················· | ···················· | NNN················· | ···················· | 1 |

| | | | | | | | | |
|---|---|---|---|---|---|---|---|---|
| G | 1853 | hCoV-19/Australia/VIC3457/2020\|EPI_ISL_521121\|2020 | ···················· | ···················· | ····················· | ···················· | ···················· | ···················· |
| G | 1854 | hCoV-19/Australia/VIC3459/2020\|EPI_ISL_521123\|2020 | ···················· | ···················· | ····················· | ···················· | ···················· | ···················· |
| G | 1855 | hCoV-19/Australia/VIC3461/2020\|EPI_ISL_521124\|2020 | ···················· | ···················· | ····················· | ···················· | ···················· | ···················· |
| G | 1857 | hCoV-19/Australia/VIC3465/2020\|EPI_ISL_521128\|2020 | ···················· | ···················· | ····················· | ···················· | ···················· | ···················· |
| G | 1858 | hCoV-19/Australia/VIC3470/2020\|EPI_ISL_521131\|2020 | ···················· | ···················· | ····················· | ···················· | ···················· | ···················· |
| G | 1859 | hCoV-19/Australia/VIC3472/2020\|EPI_ISL_521133\|2020 | ···················· | ···················· | ····················· | ···················· | ···················· | ···················· |
| G | 1861 | hCoV-19/Australia/VIC3475/2020\|EPI_ISL_521136\|2020 | ···················· | ···················· | ····················· | ···················· | ···················· | ···················· |
| G | 1862 | hCoV-19/Australia/VIC3476/2020\|EPI_ISL_521137\|2020 | ···················· | ···················· | ····················· | ···················· | ···················· | ···················· |
| G | 1863 | hCoV-19/Australia/VIC6376/2020\|EPI_ISL_521170\|2020 | ···················· | ···················· | ····················· | ···················· | ···················· | ···················· |
| G | 1864 | hCoV-19/Australia/VIC6377/2020\|EPI_ISL_521171\|2020 | ···················· | ···················· | ····················· | ···················· | ···················· | ···················· |
| G | 1865 | hCoV-19/Australia/VIC6380/2020\|EPI_ISL_521172\|2020 | ···················· | ···················· | ····················· | ···················· | ···················· | ···················· |
| G | 1866 | hCoV-19/Australia/VIC6381/2020\|EPI_ISL_521173\|2020 | ···················· | ···················· | ····················· | ···················· | ···················· | ···················· |
| G | 1867 | hCoV-19/Australia/VIC6382/2020\|EPI_ISL_521174\|2020 | ···················· | ···················· | ····················· | ···················· | ···················· | ···················· |
| G | 1868 | hCoV-19/Australia/VIC6383/2020\|EPI_ISL_521175\|2020 | ···················· | ···················· | ····················· | ···················· | ···················· | ···················· |
| G | 1869 | hCoV-19/Australia/VIC6384/2020\|EPI_ISL_521176\|2020 | ···················· | ···················· | ····················· | ···················· | ···················· | ···················· |
| G | 1870 | hCoV-19/Australia/VIC6385/2020\|EPI_ISL_521177\|2020 | ···················· | ···················· | ····················· | ···················· | ···················· | ···················· |
| G | 1871 | hCoV-19/Australia/VIC6386/2020\|EPI_ISL_521178\|2020 | ···················· | ···················· | ····················· | ···················· | ···················· | ···················· |
| G | 1872 | hCoV-19/Australia/VIC6387/2020\|EPI_ISL_521179\|2020 | ···················· | ···················· | ····················· | ···················· | ···················· | ···················· |
| G | 1873 | hCoV-19/Australia/VIC6388/2020\|EPI_ISL_521180\|2020 | ···················· | ···················· | ····················· | ···················· | ···················· | ···················· |
| G | 1874 | hCoV-19/Australia/VIC6389/2020\|EPI_ISL_521181\|2020 | ···················· | ···················· | ····················· | ···················· | ···················· | ···················· |
| G | 1875 | hCoV-19/Australia/VIC6390/2020\|EPI_ISL_521182\|2020 | ···················· | ···················· | ····················· | ···················· | ···················· | ···················· |
| G | 1876 | hCoV-19/Australia/VIC6391/2020\|EPI_ISL_521183\|2020 | ···················· | ···················· | ····················· | ···················· | ···················· | ···················· |
| G | 1877 | hCoV-19/Australia/VIC6392/2020\|EPI_ISL_521184\|2020 | ···················· | ···················· | ····················· | ···················· | ···················· | ···················· |
| G | 1878 | hCoV-19/Australia/VIC6393/2020\|EPI_ISL_521185\|2020 | ···················· | ···················· | ····················· | ···················· | ···················· | ···················· |
| G | 1879 | hCoV-19/Australia/VIC6394/2020\|EPI_ISL_521186\|2020 | ···················· | ···················· | ····················· | ···················· | ···················· | ···················· |
| G | 1880 | hCoV-19/Australia/VIC6395/2020\|EPI_ISL_521187\|2020 | ···················· | ···················· | ····················· | ···················· | ···················· | ···················· |
| G | 1881 | hCoV-19/Australia/VIC6396/2020\|EPI_ISL_521188\|2020 | ···················· | ···················· | ····················· | ···················· | ···················· | ···················· |
| G | 1882 | hCoV-19/Australia/VIC6397/2020\|EPI_ISL_521189\|2020 | ···················· | ···················· | ····················· | ···················· | ···················· | ···················· |
| G | 1883 | hCoV-19/Australia/VIC6398/2020\|EPI_ISL_521190\|2020 | ···················· | ···················· | ····················· | ···················· | ···················· | ···················· |
| G | 1884 | hCoV-19/Australia/VIC6399/2020\|EPI_ISL_521191\|2020 | ···················· | ···················· | ····················· | ···················· | ···················· | ···················· |
| G | 1885 | hCoV-19/Australia/VIC6400/2020\|EPI_ISL_521192\|2020 | ···················· | ···················· | ····················· | ···················· | ···················· | ···················· |
| G | 1886 | hCoV-19/Australia/VIC6401/2020\|EPI_ISL_521193\|2020 | ···················· | ···················· | ····················· | ···················· | ···················· | ···················· |
| G | 1887 | hCoV-19/Australia/VIC6402/2020\|EPI_ISL_521194\|2020 | ···················· | ···················· | ····················· | ···················· | ···················· | ···················· |
| G | 1888 | hCoV-19/Australia/VIC6403/2020\|EPI_ISL_521195\|2020 | ···················· | ···················· | ····················· | ···················· | ···················· | ···················· |
| G | 1889 | hCoV-19/Australia/VIC6404/2020\|EPI_ISL_521196\|2020 | ···················· | ···················· | ····················· | ···················· | ···················· | ···················· |
| G | 1890 | hCoV-19/Australia/VIC6405/2020\|EPI_ISL_521197\|2020 | ···················· | ···················· | ····················· | ···················· | ···················· | ···················· |
| G | 1891 | hCoV-19/Australia/VIC6406/2020\|EPI_ISL_521198\|2020 | ···················· | ···················· | ····················· | ···················· | ···················· | ···················· |
| G | 1892 | hCoV-19/Australia/VIC6407/2020\|EPI_ISL_521199\|2020 | ···················· | ···················· | ····················· | ···················· | ···················· | ···················· |
| G | 1893 | hCoV-19/USA/AK-PHL445/2020\|EPI_ISL_522400\|2020-06- | ···················· | ···················· | ····················· | ···················· | ···················· | ···················· |
| G | 1894 | hCoV-19/USA/AK-PHL465/2020\|EPI_ISL_522401\|2020-06- | ···················· | ···················· | ····················· | ···················· | ···················· | ···················· |
| G | 1895 | hCoV-19/USA/AK-PHL471/2020\|EPI_ISL_522402\|2020-06- | ···················· | ···················· | ····················· | ···················· | ···················· | ···················· |
| G | 1896 | hCoV-19/USA/AK-PHL529/2020\|EPI_ISL_522403\|2020-06- | ···················· | ···················· | ····················· | ···················· | ···················· | ···················· |
| G | 1897 | hCoV-19/USA/AK-PHL543/2020\|EPI_ISL_522404\|2020-06- | ···················· | ···················· | ····················· | ···················· | ···················· | ···················· |
| G | 1898 | hCoV-19/USA/AK-PHL550/2020\|EPI_ISL_522405\|2020-06- | ···················· | ·A·················· | ····················· | ···················· | ···················· | ···················· |
| G | 1899 | hCoV-19/India/MH-MW37/2020\|EPI_ISL_522437\|2020-06- | ···················· | ···················· | ····················· | ···················· | ···················· | ···················· |
| G | 1900 | hCoV-19/South Korea/KCDC2657/2020\|EPI_ISL_522440\|2 | ···················· | ·A·················· | ····················· | ···················· | ···················· | ···················· |
| G | 1901 | hCoV-19/South Korea/KCDC2658/2020\|EPI_ISL_522441\|2 | ···················· | ···················· | ····················· | ···················· | ···················· | ···················· |
| G | 1902 | hCoV-19/South Korea/KCDC2659/2020\|EPI_ISL_522442\|2 | ···················· | ···················· | ····················· | ···················· | ···················· | ···················· |
| G | 1903 | hCoV-19/South Korea/KCDC2660/2020\|EPI_ISL_522443\|2 | ···················· | ·A·················· | ····················· | ···················· | ···················· | ···················· |
| G | 1904 | hCoV-19/South Korea/KCDC2661/2020\|EPI_ISL_522444\|2 | ···················· | ·A·················· | ····················· | ···················· | ···················· | ··A················· |
| G | 1905 | hCoV-19/South Korea/KCDC2662/2020\|EPI_ISL_522445\|2 | ···················· | ···················· | ····················· | ···················· | ···················· | ···················· |
| G | 1906 | hCoV-19/South Korea/KCDC2664/2020\|EPI_ISL_522446\|2 | ···················· | ···················· | ····················· | ···················· | ···················· | ···················· |
| G | 1907 | hCoV-19/South Korea/KCDC2665/2020\|EPI_ISL_522447\|2 | ···················· | ·A·················· | ····················· | ···················· | ···················· | ···················· |
| G | 1908 | hCoV-19/South Korea/KCDC2666/2020\|EPI_ISL_522448\|2 | ···················· | ···················· | ····················· | ···················· | ···················· | ···················· |
| G | 1909 | hCoV-19/South Korea/KCDC2667/2020\|EPI_ISL_522449\|2 | ···················· | ···················· | ····················· | ···················· | ···················· | ···················· |
| G | 1910 | hCoV-19/South Korea/KCDC2668/2020\|EPI_ISL_522450\|2 | ···················· | ·A·················· | ····················· | ···················· | ···················· | ··A················· |
| G | 1911 | hCoV-19/South Korea/KCDC2669/2020\|EPI_ISL_522451\|2 | ···················· | ···················· | ····················· | ···················· | ···················· | ···················· |
| G | 1912 | hCoV-19/South Korea/KCDC2670/2020\|EPI_ISL_522452\|2 | ···················· | ···················· | ····················· | ···················· | ···················· | ···················· |
| G | 1913 | hCoV-19/South Korea/KCDC2671/2020\|EPI_ISL_522453\|2 | ···················· | ···················· | ····················· | ···················· | ···················· | ···················· |
| G | 1914 | hCoV-19/South Korea/KCDC2673/2020\|EPI_ISL_522455\|2 | ···················· | ···················· | ····················· | ···················· | ···················· | ···················· |
| G | 1915 | hCoV-19/South Korea/KCDC2674/2020\|EPI_ISL_522456\|2 | ···················· | ···················· | ····················· | ···················· | ···················· | ···················· |
| G | 1916 | hCoV-19/South Korea/KCDC2676/2020\|EPI_ISL_522458\|2 | ···················· | ···················· | ····················· | ···················· | ···················· | ···················· |
| G | 1917 | hCoV-19/South Korea/KCDC2677/2020\|EPI_ISL_522459\|2 | ···················· | ···················· | ····················· | ···················· | ···················· | ···················· |
| G | 1918 | hCoV-19/South Korea/KCDC2678/2020\|EPI_ISL_522460\|2 | ···················· | ···················· | ····················· | ···················· | ···················· | ···················· |
| G | 1919 | hCoV-19/South Korea/KCDC2679/2020\|EPI_ISL_522461\|2 | ···················· | ···················· | ····················· | ···················· | ···················· | ···················· |
| G | 1920 | hCoV-19/South Korea/KCDC2680/2020\|EPI_ISL_522462\|2 | ···················· | ···················· | ····················· | ···················· | ···················· | ···················· |
| G | 1921 | hCoV-19/South Korea/KCDC2681/2020\|EPI_ISL_522463\|2 | ···················· | ···················· | ····················· | ···················· | ···················· | ···················· |
| G | 1922 | hCoV-19/South Korea/KCDC2685/2020\|EPI_ISL_522465\|2 | ···················· | ···················· | ····················· | ···················· | ···················· | ···················· |
| G | 1923 | hCoV-19/South Korea/KCDC2686/2020\|EPI_ISL_522466\|2 | ···················· | ···················· | ····················· | ···················· | ···················· | ···················· |
| G | 1924 | hCoV-19/South Korea/KCDC2687/2020\|EPI_ISL_522467\|2 | ···················· | ···················· | ····················· | ···················· | ···················· | ···················· |
| G | 1925 | hCoV-19/South Korea/KCDC2688/2020\|EPI_ISL_522468\|2 | ···················· | ···················· | ····················· | ···················· | ···················· | ···················· |
| G | 1926 | hCoV-19/Netherlands/ZH-EMC-372/2020\|EPI_ISL_523712 | ···················· | ···················· | ····················· | ···················· | ···················· | ···················· |
| G | 1927 | hCoV-19/Netherlands/ZH-EMC-373/2020\|EPI_ISL_523713 | ···················· | ···················· | ····················· | ···················· | ···················· | ···················· |
| G | 1928 | hCoV-19/Netherlands/ZH-EMC-374/2020\|EPI_ISL_523714 | ···················· | ···················· | ····················· | ···················· | ···················· | ···················· |
| G | 1929 | hCoV-19/Netherlands/ZH-EMC-375/2020\|EPI_ISL_523715 | ···················· | ···················· | ····················· | ···················· | ···················· | ···················· |
| G | 1930 | hCoV-19/Netherlands/ZH-EMC-376/2020\|EPI_ISL_523716 | ···················· | ···················· | ····················· | ···················· | ···················· | ···················· |
| G | 1931 | hCoV-19/Netherlands/ZH-EMC-377/2020\|EPI_ISL_523717 | ···················· | ···················· | ····················· | ···················· | ···················· | ···················· |
| G | 1932 | hCoV-19/Netherlands/ZH-EMC-378/2020\|EPI_ISL_523718 | ···················· | ·A·················· | ····················· | ···················· | ···················· | ··A················· |
| G | 1933 | hCoV-19/Netherlands/ZH-EMC-379/2020\|EPI_ISL_523719 | ···················· | ···················· | ····················· | ···················· | ···················· | ···················· |
| G | 1934 | hCoV-19/Netherlands/ZH-EMC-380/2020\|EPI_ISL_523720 | ···················· | ···················· | ····················· | ···················· | ···················· | ···················· |

| | | | | | | | | |
|---|---|---|---|---|---|---|---|---|
| G | 1935 | hCoV-19/Netherlands/ZH-EMC-381/2020\|EPI_ISL_523721 | .................... | .................... | .................... | .................... | .................... | .................... |
| G | 1936 | hCoV-19/Netherlands/ZH-EMC-382/2020\|EPI_ISL_523722 | .................... | .................... | .................... | .................... | .................... | .................... |
| G | 1937 | hCoV-19/Netherlands/ZH-EMC-383/2020\|EPI_ISL_523723 | .................... | .................... | .................... | .................... | .................... | .................... |
| G | 1938 | hCoV-19/Netherlands/ZH-EMC-384/2020\|EPI_ISL_523724 | .................... | .................... | .................... | .................... | .................... | .................... |
| G | 1939 | hCoV-19/Netherlands/ZH-EMC-385/2020\|EPI_ISL_523725 | .................... | .................... | .................... | .................... | .................... | .................... |
| G | 1940 | hCoV-19/Netherlands/ZH-EMC-386/2020\|EPI_ISL_523726 | .................... | .................... | .................... | .................... | .................... | .................... |
| G | 1941 | hCoV-19/Netherlands/ZH-EMC-387/2020\|EPI_ISL_523727 | .................... | .................... | .................... | .................... | .................... | .................... |
| G | 1942 | hCoV-19/Netherlands/ZH-EMC-388/2020\|EPI_ISL_523728 | .................... | .................... | .................... | .................... | .................... | .................... |
| G | 1943 | hCoV-19/Netherlands/ZH-EMC-389/2020\|EPI_ISL_523729 | .................... | .................... | .................... | .................... | .................... | .................... |
| G | 1944 | hCoV-19/Netherlands/ZH-EMC-390/2020\|EPI_ISL_523730 | .................... | .................... | .................... | .................... | .................... | .................... |
| G | 1945 | hCoV-19/Netherlands/ZH-EMC-391/2020\|EPI_ISL_523731 | .................... | .................... | .................... | .................... | .................... | .................... |
| G | 1946 | hCoV-19/Netherlands/ZH-EMC-392/2020\|EPI_ISL_523732 | .................... | .................... | .................... | .................... | .................... | .................... |
| G | 1947 | hCoV-19/Netherlands/ZH-EMC-393/2020\|EPI_ISL_523733 | .................... | .................... | .................... | .................... | .................... | .................... |
| G | 1948 | hCoV-19/Netherlands/ZH-EMC-394/2020\|EPI_ISL_523734 | .................... | .................... | .................... | .................... | .................... | .................... |
| G | 1949 | hCoV-19/Netherlands/ZH-EMC-395/2020\|EPI_ISL_523735 | .................... | .................... | .................... | .................... | .................... | .................... |
| G | 1950 | hCoV-19/Netherlands/ZH-EMC-396/2020\|EPI_ISL_523736 | .................... | .................... | .................... | .................... | .................... | .................... |
| G | 1951 | hCoV-19/Netherlands/ZH-EMC-397/2020\|EPI_ISL_523737 | .................... | .................... | .................... | .................... | .................... | .................... |
| G | 1952 | hCoV-19/Netherlands/ZH-EMC-398/2020\|EPI_ISL_523738 | .................... | .................... | .................... | .................... | .................... | .................... |
| G | 1953 | hCoV-19/Netherlands/ZH-EMC-399/2020\|EPI_ISL_523739 | .................... | .................... | .................... | .................... | .................... | .................... |
| G | 1954 | hCoV-19/Netherlands/ZH-EMC-400/2020\|EPI_ISL_523740 | .................... | .................... | .................... | .................... | .................... | .................... |
| G | 1955 | hCoV-19/Netherlands/ZH-EMC-401/2020\|EPI_ISL_523741 | .................... | .................... | .................... | .................... | .................... | .................... |
| G | 1956 | hCoV-19/Netherlands/ZH-EMC-402/2020\|EPI_ISL_523742 | .................... | .................... | .................... | .................... | .................... | .................... |
| G | 1957 | hCoV-19/Netherlands/ZH-EMC-404/2020\|EPI_ISL_523744 | .................... | .................... | .................... | .................... | .................... | .................... |
| G | 1958 | hCoV-19/Netherlands/ZH-EMC-405/2020\|EPI_ISL_523745 | .................... | .................... | .................... | .................... | .................... | .................... |
| G | 1959 | hCoV-19/Netherlands/ZH-EMC-406/2020\|EPI_ISL_523746 | .................... | .................... | .................... | .................... | .................... | .................... |
| G | 1960 | hCoV-19/Netherlands/ZH-EMC-407/2020\|EPI_ISL_523747 | .................... | .................... | .................... | .................... | .................... | .................... |
| G | 1961 | hCoV-19/Netherlands/ZH-EMC-408/2020\|EPI_ISL_523748 | .................... | .................... | .................... | .................... | .................... | .................... |
| G | 1962 | hCoV-19/Netherlands/ZH-EMC-409/2020\|EPI_ISL_523749 | .................... | .................... | .................... | .................... | .................... | .................... |
| G | 1963 | hCoV-19/Netherlands/ZH-EMC-410/2020\|EPI_ISL_523750 | .................... | .................... | .................... | .................... | .................... | .................... |
| G | 1964 | hCoV-19/Netherlands/ZH-EMC-411/2020\|EPI_ISL_523751 | .................... | .................... | .................... | .................... | .................... | .................... |
| G | 1965 | hCoV-19/Netherlands/ZH-EMC-412/2020\|EPI_ISL_523752 | .................... | .................... | .................... | .................... | .................... | .................... |
| G | 1966 | hCoV-19/Netherlands/ZH-EMC-413/2020\|EPI_ISL_523753 | .................... | .................... | .................... | .................... | .................... | .................... |
| G | 1967 | hCoV-19/Netherlands/ZH-EMC-414/2020\|EPI_ISL_523754 | .................... | .................... | .................... | .................... | .................... | .................... |
| G | 1968 | hCoV-19/Netherlands/ZH-EMC-415/2020\|EPI_ISL_523755 | .................... | .................... | .................... | .................... | .................... | .................... |
| G | 1969 | hCoV-19/Netherlands/ZH-EMC-416/2020\|EPI_ISL_523756 | .................... | .................... | .................... | .................... | .................... | .................... |
| G | 1970 | hCoV-19/Netherlands/ZH-EMC-417/2020\|EPI_ISL_523757 | .................... | .................... | .................... | .................... | .................... | .................... |
| G | 1971 | hCoV-19/Netherlands/ZH-EMC-418/2020\|EPI_ISL_523758 | .................... | .................... | .................... | .................... | .................... | .................... |
| G | 1972 | hCoV-19/Netherlands/ZH-EMC-419/2020\|EPI_ISL_523759 | .................... | .................... | .................... | .................... | .................... | .................... |
| G | 1973 | hCoV-19/Netherlands/ZH-EMC-421/2020\|EPI_ISL_523760 | .................... | .................... | .................... | .................... | .................... | .................... |
| G | 1974 | hCoV-19/Netherlands/ZH-EMC-422/2020\|EPI_ISL_523761 | .................... | .................... | .................... | .................... | .................... | .................... |
| G | 1975 | hCoV-19/Netherlands/ZH-EMC-423/2020\|EPI_ISL_523762 | .................... | .................... | .................... | .................... | .................... | .................... |
| G | 1976 | hCoV-19/Netherlands/ZH-EMC-424/2020\|EPI_ISL_523763 | .................... | .................... | .................... | .................... | .................... | .................... |
| G | 1977 | hCoV-19/Netherlands/ZH-EMC-425/2020\|EPI_ISL_523764 | .................... | .................... | .................... | .................... | .................... | .................... |
| G | 1978 | hCoV-19/Netherlands/ZH-EMC-426/2020\|EPI_ISL_523765 | .................... | .................... | .................... | .................... | .................... | .................... |
| G | 1979 | hCoV-19/Netherlands/ZH-EMC-427/2020\|EPI_ISL_523766 | .................... | .................... | .................... | .................... | .................... | .................... |
| G | 1980 | hCoV-19/Netherlands/ZH-EMC-429/2020\|EPI_ISL_523767 | .................... | .................... | .................... | .................... | .................... | .................... |
| G | 1981 | hCoV-19/Netherlands/ZH-EMC-430/2020\|EPI_ISL_523768 | .................... | .................... | .................... | .................... | .................... | .................... |
| G | 1982 | hCoV-19/Netherlands/ZH-EMC-431/2020\|EPI_ISL_523769 | .................... | .................... | .................... | .................... | .................... | .................... |
| G | 1983 | hCoV-19/Netherlands/ZH-EMC-432/2020\|EPI_ISL_523770 | .................... | .................... | .................... | .................... | .................... | .................... |
| G | 1984 | hCoV-19/Netherlands/ZH-EMC-433/2020\|EPI_ISL_523771 | .................... | .................... | .................... | .................... | .................... | .................... |
| G | 1985 | hCoV-19/Netherlands/ZH-EMC-434/2020\|EPI_ISL_523772 | .................... | .................... | .................... | .................... | .................... | .................... |
| G | 1986 | hCoV-19/Netherlands/ZH-EMC-435/2020\|EPI_ISL_523773 | .................... | .................... | .................... | .................... | .................... | .................... |
| G | 1987 | hCoV-19/Netherlands/ZH-EMC-436/2020\|EPI_ISL_523774 | .................... | .................... | .................... | .................... | .................... | .................... |
| G | 1988 | hCoV-19/Netherlands/ZH-EMC-437/2020\|EPI_ISL_523775 | .................... | .................... | .................... | .................... | .................... | .................... |
| G | 1989 | hCoV-19/Netherlands/ZH-EMC-438/2020\|EPI_ISL_523776 | .................... | .................... | .................... | .................... | .................... | .................... |
| G | 1990 | hCoV-19/Netherlands/ZH-EMC-439/2020\|EPI_ISL_523777 | .................... | .................... | .................... | .................... | .................... | .................... |
| G | 1991 | hCoV-19/Netherlands/ZH-EMC-440/2020\|EPI_ISL_523778 | .................... | .................... | .................... | .................... | .................... | .................... |
| G | 1992 | hCoV-19/Netherlands/ZH-EMC-441/2020\|EPI_ISL_523779 | .................... | .................... | .................... | .................... | .................... | .................... |
| G | 1993 | hCoV-19/Netherlands/ZH-EMC-442/2020\|EPI_ISL_523780 | .................... | .................... | .................... | .................... | .................... | .................... |
| G | 1994 | hCoV-19/Netherlands/ZH-EMC-444/2020\|EPI_ISL_523782 | .................... | .................... | .................... | .................... | .................... | .................... |
| G | 1995 | hCoV-19/Netherlands/ZH-EMC-445/2020\|EPI_ISL_523783 | .................... | .................... | .................... | .................... | .................... | .................... |
| G | 1996 | hCoV-19/Netherlands/ZH-EMC-446/2020\|EPI_ISL_523784 | .................... | .................... | .................... | .................... | .................... | .................... |
| G | 1997 | hCoV-19/Netherlands/ZH-EMC-447/2020\|EPI_ISL_523785 | .................... | .................... | .................... | .................... | .................... | .................... |
| G | 1998 | hCoV-19/Netherlands/ZH-EMC-448/2020\|EPI_ISL_523786 | .................... | .................... | .................... | .................... | .................... | .................... |
| G | 1999 | hCoV-19/Netherlands/ZH-EMC-449/2020\|EPI_ISL_523787 | .................... | .................... | .................... | .................... | .................... | .................... |
| G | 2000 | hCoV-19/Netherlands/ZH-EMC-450/2020\|EPI_ISL_523788 | .................... | .................... | .................... | .................... | .................... | .................... |
| G | 2001 | hCoV-19/Netherlands/ZH-EMC-451/2020\|EPI_ISL_523789 | .................... | .................... | .................... | .................... | .................... | .................... |
| G | 2002 | hCoV-19/Netherlands/ZH-EMC-452/2020\|EPI_ISL_523790 | .................... | .................... | .................... | .................... | .................... | .................... |
| G | 2003 | hCoV-19/Netherlands/ZH-EMC-453/2020\|EPI_ISL_523791 | .................... | .................... | .................... | .................... | .................... | .................... |
| G | 2004 | hCoV-19/Netherlands/ZH-EMC-454/2020\|EPI_ISL_523792 | .................... | .................... | .................... | .................... | .................... | .................... |
| G | 2005 | hCoV-19/Netherlands/ZH-EMC-455/2020\|EPI_ISL_523793 | .................... | .................... | .................... | .................... | .................... | .................... |
| G | 2006 | hCoV-19/Netherlands/ZH-EMC-456/2020\|EPI_ISL_523794 | .................... | .................... | .................... | .................... | .................... | .................... |
| G | 2007 | hCoV-19/Netherlands/ZH-EMC-457/2020\|EPI_ISL_523795 | .................... | .................... | .................... | .................... | .................... | .................... |
| G | 2008 | hCoV-19/Netherlands/ZH-EMC-459/2020\|EPI_ISL_523796 | .................... | .................... | .................... | .................... | .................... | .................... |
| G | 2009 | hCoV-19/Netherlands/ZH-EMC-460/2020\|EPI_ISL_523797 | .................... | .................... | .................... | .................... | .................... | .................... |
| G | 2010 | hCoV-19/Netherlands/ZH-EMC-461/2020\|EPI_ISL_523798 | .................... | .................... | .................... | .................... | .................... | .................... |
| G | 2011 | hCoV-19/Netherlands/ZH-EMC-462/2020\|EPI_ISL_523799 | .................... | .................... | .................... | .................... | .................... | .................... |
| G | 2012 | hCoV-19/Australia/NSW809/2020\|EPI_ISL_526151\|2020- | .................... | .................... | .................... | .................... | .................... | .................... |
| G | 2013 | hCoV-19/Australia/NSW810/2020\|EPI_ISL_526152\|2020- | .................... | .................... | .................... | .................... | .................... | .................... |
| G | 2014 | hCoV-19/Australia/NSW815/2020\|EPI_ISL_526157\|2020- | .................... | .................... | .................... | .................... | .................... | .................... |

| | | | | | | | |
|---|---|---|---|---|---|---|---|
| G | 2015 hCoV-19/Australia/NSW816/2020\|EPI_ISL_526158\|2020- | ···················· | ···················· | ···················· | ···················· | ···················· | ···················· |
| G | 2016 hCoV-19/Australia/NSW817/2020\|EPI_ISL_526159\|2020- | ···················· | ···················· | ···················· | ···················· | ···················· | ···················· |
| G | 2017 hCoV-19/USA/CO-CSU-E0491/2020\|EPI_ISL_527482\|2020- | ···················· | ···················· | ···················· | ···················· | ···················· | ···················· |
| G | 2018 hCoV-19/USA/CO-CSU-E0983-1/2020\|EPI_ISL_527483\|202 | ···················· | ···················· | ···················· | ···················· | ···················· | ···················· |
| G | 2019 hCoV-19/USA/CO-CSU-E2959/2020\|EPI_ISL_527488\|2020- | ···················· | ···················· | ···················· | ···················· | ···················· | ···················· |
| G | 2020 hCoV-19/DRC/10371/2020\|EPI_ISL_527489\|2020-05-22 | ···················· | ···················· | ···················· | ···················· | ···················· | ···················· |
| G | 2021 hCoV-19/USA/VA-DCLS-1096/2020\|EPI_ISL_529908\|2020- | ···················· | ···················· | ···················· | ···················· | ···················· | ···················· |
| G | 2022 hCoV-19/USA/VA-DCLS-1102/2020\|EPI_ISL_529909\|2020- | ···················· | ·A·················· | ···················· | ···················· | ···················· | ···················· |
| G | 2023 hCoV-19/USA/VA-DCLS-1103/2020\|EPI_ISL_529910\|2020- | ···················· | ·A·················· | ···················· | ···················· | ···················· | ···················· |
| G | 2024 hCoV-19/USA/VA-DCLS-1108/2020\|EPI_ISL_529912\|2020- | ···················· | ···················· | ···················· | ···················· | ···················· | ···················· |
| G | 2025 hCoV-19/USA/VA-DCLS-1110/2020\|EPI_ISL_529913\|2020- | ···················· | ···················· | ···················· | ···················· | ···················· | ···················· |
| G | 2026 hCoV-19/USA/VA-DCLS-1111/2020\|EPI_ISL_529914\|2020- | ···················· | ···················· | ···················· | ···················· | ···················· | ···················· |
| G | 2027 hCoV-19/USA/VA-DCLS-1113/2020\|EPI_ISL_529915\|2020- | ···················· | ···················· | ···················· | ···················· | ···················· | ···················· |
| G | 2028 hCoV-19/USA/VA-DCLS-1114/2020\|EPI_ISL_529916\|2020- | ···················· | ···················· | ···················· | ···················· | ···················· | ·········A·········· |
| G | 2029 hCoV-19/USA/VA-DCLS-1116/2020\|EPI_ISL_529917\|2020- | ···················· | ···················· | ···················· | ···················· | ···················· | ···················· |
| G | 2030 hCoV-19/USA/VA-DCLS-1120/2020\|EPI_ISL_529918\|2020- | ···················· | ···················· | ···················· | ···················· | ···················· | ···················· |
| G | 2031 hCoV-19/USA/VA-DCLS-1122/2020\|EPI_ISL_529919\|2020- | ···················· | ···················· | ···················· | ···················· | ···················· | ···················· |
| G | 2032 hCoV-19/USA/VA-DCLS-1123/2020\|EPI_ISL_529920\|2020- | ···················· | ···················· | ···················· | ···················· | ···················· | ···················· |
| G | 2033 hCoV-19/USA/VA-DCLS-1124/2020\|EPI_ISL_529921\|2020- | ···················· | ···················· | ···················· | ···················· | ···················· | ···················· |
| G | 2034 hCoV-19/USA/VA-DCLS-1130/2020\|EPI_ISL_529923\|2020- | ···················· | ···················· | ···················· | ···················· | ···················· | ···················· |
| G | 2035 hCoV-19/USA/VA-DCLS-1131/2020\|EPI_ISL_529924\|2020- | ···················· | ···················· | ···················· | ···················· | ···················· | ···················· |
| G | 2036 hCoV-19/USA/VA-DCLS-1133/2020\|EPI_ISL_529925\|2020- | ···················· | ···················· | ···················· | ···················· | ···················· | ···················· |
| G | 2037 hCoV-19/USA/VA-DCLS-1134/2020\|EPI_ISL_529926\|2020- | ···················· | ···················· | ···················· | ···················· | ···················· | ···················· |
| G | 2038 hCoV-19/USA/VA-DCLS-1137/2020\|EPI_ISL_529927\|2020- | ···················· | ···················· | ···················· | ···················· | ···················· | ···················· |
| G | 2039 hCoV-19/USA/VA-DCLS-1138/2020\|EPI_ISL_529928\|2020- | ···················· | ···················· | ···················· | ···················· | ···················· | ···················· |
| G | 2040 hCoV-19/USA/VA-DCLS-1139/2020\|EPI_ISL_529929\|2020- | ···················· | ···················· | ···················· | ···················· | ···················· | ···················· |
| G | 2041 hCoV-19/USA/VA-DCLS-1141/2020\|EPI_ISL_529930\|2020- | ···················· | ···················· | ···················· | ···················· | ···················· | ···················· |
| G | 2042 hCoV-19/USA/VA-DCLS-1142/2020\|EPI_ISL_529931\|2020- | ···················· | ···················· | ···················· | ···················· | ···················· | ···················· |
| G | 2043 hCoV-19/USA/VA-DCLS-1144/2020\|EPI_ISL_529932\|2020- | ···················· | ···················· | ···················· | ···················· | ···················· | ···················· |
| G | 2044 hCoV-19/USA/VA-DCLS-1145/2020\|EPI_ISL_529933\|2020- | ···················· | ···················· | ···················· | ···················· | ···················· | ···················· |
| G | 2045 hCoV-19/USA/VA-DCLS-1147/2020\|EPI_ISL_529934\|2020- | ···················· | ···················· | ···················· | ···················· | ···················· | ···················· |
| G | 2046 hCoV-19/USA/VA-DCLS-1149/2020\|EPI_ISL_529935\|2020- | ···················· | ···················· | ···················· | ···················· | ···················· | ···················· |
| G | 2047 hCoV-19/USA/VA-DCLS-1189/2020\|EPI_ISL_529936\|2020- | ···················· | ···················· | ···················· | ···················· | ···················· | ···················· |
| G | 2048 hCoV-19/USA/VA-DCLS-1244/2020\|EPI_ISL_529937\|2020- | ···················· | ···················· | ···················· | ···················· | ···················· | ···················· |
| G | 2049 hCoV-19/USA/FL-BPHL-1010/2020\|EPI_ISL_517900\|2020- | ···················· | ···················· | ···················· | ···················· | ···················· | ···················· |
| G | 2051 hCoV-19/USA/FL-BPHL-1013/2020\|EPI_ISL_517903\|2020- | ···················· | ···················· | ···················· | ···················· | ···················· | ···················· |
| G | 2053 hCoV-19/USA/FL-BPHL-1021/2020\|EPI_ISL_517911\|2020- | ···················· | ···················· | ···················· | ···················· | ···················· | ···················· |
| G | 2054 hCoV-19/Peru/LIM-INS-048/2020\|EPI_ISL_517959\|2020- | ···················· | ···················· | ···················· | ···················· | ···················· | ···················· |
| G | 2055 hCoV-19/USA/TX-DSHS-0036/2020\|EPI_ISL_517969\|2020- | ···················· | ···················· | ···················· | ···················· | ···················· | ···················· |
| G | 2056 hCoV-19/Singapore/766_2/2020\|EPI_ISL_517996\|2020-0 | ···················· | ···················· | ···················· | ···················· | ···················· | ···················· |
| G | 2057 hCoV-19/Singapore/834/2020\|EPI_ISL_517997\|2020-06- | ···················· | ···················· | ···················· | ···················· | ···················· | ···················· |
| G | 2058 hCoV-19/Singapore/835/2020\|EPI_ISL_517998\|2020-06- | ···················· | ···················· | ···················· | ···················· | ···················· | ···················· |
| G | 2059 hCoV-19/Singapore/836/2020\|EPI_ISL_517999\|2020-06- | ···················· | ···················· | ···················· | ···················· | ···················· | ···················· |
| G | 2061 hCoV-19/Australia/VIC3478/2020\|EPI_ISL_521139\|2020 | ···················· | ···················· | ···················· | ···················· | ··NN················ | ···················· |
| G | 2062 hCoV-19/Australia/VIC3479/2020\|EPI_ISL_521140\|2020 | ···················· | ···················· | ···················· | ···················· | ···················· | ···················· |
| G | 2063 hCoV-19/Australia/VIC3480/2020\|EPI_ISL_521141\|2020 | ···················· | ···················· | ···················· | ···················· | ···················· | ···················· |
| G | 2065 hCoV-19/Australia/VIC3482/2020\|EPI_ISL_521143\|2020 | ···················· | ···················· | ···················· | ···················· | ···················· | ···················· |
| G | 2066 hCoV-19/Australia/VIC3483/2020\|EPI_ISL_521144\|2020 | ···················· | ···················· | ···················· | ···················· | ···················· | ···················· |
| G | 2069 hCoV-19/Australia/VIC3486/2020\|EPI_ISL_521147\|2020 | ···················· | ···················· | ···················· | ···················· | ···················· | ···················· |
| G | 2070 hCoV-19/Australia/VIC3487/2020\|EPI_ISL_521148\|2020 | ···················· | ···················· | ···················· | ···················· | ···················· | ···················· |
| G | 2071 hCoV-19/Australia/VIC3490/2020\|EPI_ISL_521150\|2020 | ···················· | ···················· | ···················· | ···················· | ···················· | ···················· |
| G | 2072 hCoV-19/Australia/VIC3491/2020\|EPI_ISL_521151\|2020 | ···················· | ···················· | ···················· | ···················· | ···················· | ···················· |
| G | 2074 hCoV-19/USA/MN-MDH-1486/2020\|EPI_ISL_521153\|2020-0 | ···················· | ···················· | ···················· | ···················· | ···················· | ···················· |
| G | 2075 hCoV-19/Australia/VIC6358/2020\|EPI_ISL_521154\|2020 | ···················· | ···················· | ···················· | ···················· | ···················· | ···················· |
| G | 2076 hCoV-19/Australia/VIC6360/2020\|EPI_ISL_521156\|2020 | ···················· | ···················· | ···················· | ···················· | ···················· | ···················· |
| G | 2077 hCoV-19/Australia/VIC6361/2020\|EPI_ISL_521157\|2020 | ···················· | ···················· | ···················· | ···················· | ···················· | ···················· |
| G | 2078 hCoV-19/Australia/VIC6362/2020\|EPI_ISL_521158\|2020 | ···················· | ···················· | ···················· | ···················· | ···················· | ···················· |
| G | 2079 hCoV-19/Australia/VIC6363/2020\|EPI_ISL_521159\|2020 | ···················· | ···················· | ···················· | ···················· | ···················· | ···················· |
| G | 2080 hCoV-19/Australia/VIC6365/2020\|EPI_ISL_521160\|2020 | ···················· | ···················· | ···················· | ···················· | ···················· | ···················· |
| G | 2081 hCoV-19/Australia/VIC6366/2020\|EPI_ISL_521161\|2020 | ···················· | ···················· | ···················· | ···················· | ···················· | ···················· |
| G | 2082 hCoV-19/Australia/VIC6367/2020\|EPI_ISL_521162\|2020 | ···················· | ···················· | ···················· | ···················· | ···················· | ···················· |
| G | 2083 hCoV-19/Australia/VIC6368/2020\|EPI_ISL_521163\|2020 | ···················· | ···················· | ···················· | ···················· | ···················· | ···················· |
| G | 2084 hCoV-19/Australia/VIC6371/2020\|EPI_ISL_521165\|2020 | ···················· | ···················· | ···················· | ···················· | ···················· | ···················· |
| G | 2085 hCoV-19/Australia/VIC6372/2020\|EPI_ISL_521166\|2020 | ···················· | ···················· | ···················· | ···················· | ···················· | ···················· |
| G | 2086 hCoV-19/Australia/VIC6373/2020\|EPI_ISL_521167\|2020 | ···················· | ···················· | ···················· | ···················· | ···················· | ···················· |
| G | 2087 hCoV-19/Australia/VIC6374/2020\|EPI_ISL_521168\|2020 | ···················· | ···················· | ···················· | ···················· | ···················· | ···················· |
| G | 2088 hCoV-19/South Korea/KCDC2689/2020\|EPI_ISL_522469\|2 | ···················· | ···················· | ···················· | ···················· | ···················· | ···················· |
| G | 2089 hCoV-19/South Korea/KCDC2690/2020\|EPI_ISL_522470\|2 | ···················· | ···················· | ···················· | ···················· | ···················· | ···················· |
| G | 2090 hCoV-19/South Korea/KCDC2692/2020\|EPI_ISL_522472\|2 | ···················· | ···················· | ···················· | ···················· | ···················· | ···················· |
| G | 2091 hCoV-19/South Korea/KCDC2693/2020\|EPI_ISL_522473\|2 | ···················· | ···················· | ···················· | ···················· | ···················· | ···················· |
| G | 2092 hCoV-19/South Korea/KCDC2694/2020\|EPI_ISL_522474\|2 | ···················· | ···················· | ···················· | ···················· | ···················· | ···················· |
| G | 2093 hCoV-19/South Korea/KCDC2695/2020\|EPI_ISL_522475\|2 | ···················· | ···················· | ···················· | ···················· | ···················· | ···················· |
| G | 2094 hCoV-19/South Korea/KCDC2696/2020\|EPI_ISL_522476\|2 | ···················· | ···················· | ···················· | ···················· | ···················· | ···················· |
| G | 2095 hCoV-19/South Korea/KCDC2697/2020\|EPI_ISL_522477\|2 | ···················· | ···················· | ···················· | ···················· | ···················· | ···················· |
| G | 2096 hCoV-19/South Korea/KCDC2698/2020\|EPI_ISL_522478\|2 | ···················· | ···················· | ···················· | ···················· | ···················· | ···················· |
| G | 2097 hCoV-19/South Korea/KCDC2699/2020\|EPI_ISL_522479\|2 | ···················· | ···················· | ···················· | ···················· | ···················· | ···················· |
| G | 2098 hCoV-19/South Korea/KCDC2700/2020\|EPI_ISL_522480\|2 | ···················· | ···················· | ···················· | ···················· | ···················· | ···················· |
| G | 2099 hCoV-19/South Korea/KCDC2701/2020\|EPI_ISL_522481\|2 | ···················· | ···················· | ···················· | ···················· | ···················· | ···················· |
| G | 2100 hCoV-19/South Korea/KCDC2702/2020\|EPI_ISL_522482\|2 | ···················· | ···················· | ···················· | ···················· | ···················· | ···················· |
| G | 2101 hCoV-19/South Korea/KCDC2703/2020\|EPI_ISL_522483\|2 | ···················· | ···················· | ···················· | ···················· | ···················· | ···················· |

| | | | | | | | |
|---|---|---|---|---|---|---|---|
| G | 2102 hCoV-19/South Korea/KCDC2704/2020\|EPI_ISL_522484\|2 | ···················· | ···················· | ···················· | ···················· | ···················· | ···················· |
| G | 2103 hCoV-19/South Korea/KCDC2705/2020\|EPI_ISL_522485\|2 | ···················· | ···················· | ···················· | ···················· | ···················· | ···················· |
| G | 2104 hCoV-19/South Korea/KCDC2707/2020\|EPI_ISL_522486\|2 | ···················· | ···················· | ···················· | ···················· | ···················· | ···················· |
| G | 2105 hCoV-19/South Korea/KCDC2708/2020\|EPI_ISL_522487\|2 | ···················· | ···················· | ···················· | ···················· | ···················· | ···················· |
| G | 2106 hCoV-19/South Korea/KCDC2709/2020\|EPI_ISL_522488\|2 | ···················· | ···················· | ···················· | ···················· | ···················· | ···················· |
| G | 2107 hCoV-19/South Korea/KCDC2710/2020\|EPI_ISL_522489\|2 | ···················· | ···················· | ···················· | ···················· | ···················· | ···················· |
| G | 2108 hCoV-19/South Korea/KCDC2711/2020\|EPI_ISL_522490\|2 | ···················· | ···················· | ···················· | ···················· | ···················· | ···················· |
| G | 2109 hCoV-19/South Korea/KCDC2712/2020\|EPI_ISL_522491\|2 | ···················· | ···················· | ···················· | ···················· | ···················· | ···················· |
| G | 2110 hCoV-19/South Korea/KCDC2713/2020\|EPI_ISL_522492\|2 | ···················· | ···················· | ···················· | ···················· | ···················· | ···················· |
| G | 2111 hCoV-19/South Korea/KCDC2714/2020\|EPI_ISL_522493\|2 | ···················· | ···················· | ···················· | ···················· | ···················· | ···················· |
| G | 2112 hCoV-19/South Korea/KCDC2716/2020\|EPI_ISL_522495\|2 | ···················· | ···················· | ···················· | ···················· | ···················· | ···················· |
| G | 2113 hCoV-19/South Korea/KCDC2717/2020\|EPI_ISL_522496\|2 | ···················· | ···················· | ···················· | ···················· | ···················· | ···················· |
| G | 2114 hCoV-19/South Korea/KCDC2719/2020\|EPI_ISL_522498\|2 | ···················· | ···················· | ···················· | ···················· | ···················· | ···················· |
| G | 2115 hCoV-19/South Korea/KCDC2720/2020\|EPI_ISL_522499\|2 | ···················· | ···················· | ···················· | ···················· | ···················· | ···················· |
| G | 2116 hCoV-19/South Korea/KCDC2721/2020\|EPI_ISL_522500\|2 | ···················· | ···················· | ···················· | ···················· | ···················· | ···················· |
| G | 2117 hCoV-19/South Korea/KCDC2722/2020\|EPI_ISL_522501\|2 | ···················· | ···················· | ···················· | ···················· | ···················· | ···················· |
| G | 2118 hCoV-19/Dominican Republic/ICGEB_UNIBE051/2020\|EPI | ···················· | ···················· | ···················· | ···················· | ···················· | ···················· |
| G | 2119 hCoV-19/USA/OR-OHSU-1677/2020\|EPI_ISL_526000\|2020- | ···················· | ·A·················· | ···················· | ···················· | ···················· | ···················· |
| G | 2120 hCoV-19/USA/OR-OHSU-1679/2020\|EPI_ISL_526001\|2020- | ···················· | ···················· | ···················· | ···················· | ···················· | ···················· |
| G | 2121 hCoV-19/USA/OR-OHSU-1681/2020\|EPI_ISL_526002\|2020- | ···················· | ·A·················· | ···················· | ···················· | ···················· | ···················· |
| G | 2122 hCoV-19/USA/OR-OHSU-1687/2020\|EPI_ISL_526003\|2020- | ···················· | ···················· | ···················· | ···················· | ···················· | ···················· |
| G | 2123 hCoV-19/USA/OR-OHSU-1689/2020\|EPI_ISL_526004\|2020- | ···················· | ·A·················· | ···················· | ···················· | ···················· | ···················· |
| G | 2124 hCoV-19/USA/OR-OHSU-1697/2020\|EPI_ISL_526005\|2020- | ···················· | ···················· | ···················· | ···················· | ···················· | ···················· |
| G | 2125 hCoV-19/USA/OR-OHSU-1699/2020\|EPI_ISL_526006\|2020- | ···················· | ···················· | ···················· | ···················· | ···················· | ···················· |
| G | 2126 hCoV-19/USA/OR-OHSU-1701/2020\|EPI_ISL_526007\|2020- | ···················· | ···················· | ···················· | ···················· | ···················· | ···················· |
| G | 2127 hCoV-19/USA/OR-OHSU-1703/2020\|EPI_ISL_526008\|2020- | ···················· | ···················· | ···················· | ···················· | ···················· | ···················· |
| G | 2128 hCoV-19/USA/OR-OHSU-1705/2020\|EPI_ISL_526009\|2020- | ···················· | ·A·················· | ···················· | ···················· | ···················· | ···················· |
| G | 2129 hCoV-19/USA/OR-OHSU-1707/2020\|EPI_ISL_526010\|2020- | ···················· | ·A·················· | ···················· | ···················· | ···················· | ···················· |
| G | 2130 hCoV-19/USA/OR-OHSU-1711/2020\|EPI_ISL_526011\|2020- | ···················· | ·A·················· | ···················· | ···················· | ···················· | ···················· |
| G | 2131 hCoV-19/USA/OR-OHSU-1717/2020\|EPI_ISL_526012\|2020- | ···················· | ···················· | ···················· | ···················· | ···················· | ···················· |
| G | 2132 hCoV-19/USA/OR-OHSU-1719/2020\|EPI_ISL_526013\|2020- | ···················· | ···················· | ···················· | ···················· | ···················· | ···················· |
| G | 2133 hCoV-19/USA/OR-OHSU-1723/2020\|EPI_ISL_526014\|2020- | ···················· | ·A·················· | ···················· | ···················· | ···················· | ···················· |
| G | 2134 hCoV-19/USA/OR-OHSU-1729/2020\|EPI_ISL_526015\|2020- | ···················· | ···················· | ···················· | ···················· | ···················· | ···················· |
| G | 2135 hCoV-19/USA/OR-OHSU-1731/2020\|EPI_ISL_526016\|2020- | ···················· | ···················· | ···················· | ···················· | ···················· | ···················· |
| G | 2136 hCoV-19/USA/OR-OHSU-1735/2020\|EPI_ISL_526017\|2020- | ···················· | ···················· | ···················· | ···················· | ···················· | N··················· |
| G | 2137 hCoV-19/USA/OR-OHSU-1739/2020\|EPI_ISL_526018\|2020- | ···················· | ···················· | ···················· | ···················· | ···················· | ···················· |
| G | 2138 hCoV-19/USA/OR-OHSU-1747/2020\|EPI_ISL_526019\|2020- | ···················· | ·A·················· | ···················· | ···················· | ···················· | ···················· |
| G | 2139 hCoV-19/USA/OR-OHSU-1753/2020\|EPI_ISL_526020\|2020- | ···················· | ·A·················· | ···················· | ···················· | ···················· | ···················· |
| G | 2140 hCoV-19/USA/OR-OHSU-1757/2020\|EPI_ISL_526021\|2020- | ···················· | ·A·················· | ···················· | ···················· | ···················· | ···················· |
| G | 2141 hCoV-19/USA/OR-OHSU-1763/2020\|EPI_ISL_526022\|2020- | ···················· | ·A·················· | ···················· | ···················· | ···················· | ···················· |
| G | 2142 hCoV-19/USA/OR-OHSU-1773/2020\|EPI_ISL_526024\|2020- | ···················· | ···················· | ···················· | ···················· | ···················· | ···················· |
| G | 2143 hCoV-19/USA/OR-OHSU-1777/2020\|EPI_ISL_526025\|2020- | ···················· | ···················· | ···················· | ···················· | ···················· | ···················· |
| G | 2144 hCoV-19/USA/OR-OHSU-1779/2020\|EPI_ISL_526026\|2020- | ···················· | ·A·················· | ···················· | ···················· | ···················· | ···················· |
| G | 2145 hCoV-19/USA/OR-OHSU-1785/2020\|EPI_ISL_526027\|2020- | ···················· | ···················· | ···················· | ···················· | ···················· | ···················· |
| G | 2146 hCoV-19/USA/OR-OHSU-1789/2020\|EPI_ISL_526028\|2020- | ···················· | ·A·················· | ···················· | ···················· | ···················· | ···················· |
| G | 2147 hCoV-19/USA/OR-OHSU-1791/2020\|EPI_ISL_526029\|2020- | ···················· | ·A·················· | ···················· | ···················· | ···················· | ···················· |
| G | 2148 hCoV-19/USA/OR-OHSU-1985/2020\|EPI_ISL_526083\|2020- | ···················· | ···················· | ···················· | ···················· | ···················· | ···················· |
| G | 2149 hCoV-19/USA/OR-OHSU-1989/2020\|EPI_ISL_526084\|2020- | ···················· | ···················· | ···················· | ···················· | ···················· | ···················· |
| G | 2150 hCoV-19/USA/TX-OHSU-1633/2020\|EPI_ISL_526086\|2020- | ···················· | ···················· | ···················· | ···················· | ···················· | ···················· |
| G | 2151 hCoV-19/USA/WA-OHSU-0444/2020\|EPI_ISL_526087\|2020- | ···················· | ···················· | ···················· | ···················· | ···················· | ···················· |
| G | 2152 hCoV-19/USA/WA-OHSU-0462/2020\|EPI_ISL_526088\|2020- | ···················· | ···················· | ···················· | ···················· | ···················· | ···················· |
| G | 2153 hCoV-19/USA/WA-OHSU-0464/2020\|EPI_ISL_526089\|2020- | ···················· | ···················· | ···················· | ···················· | ···················· | ···················· |
| G | 2154 hCoV-19/USA/WA-OHSU-0505/2020\|EPI_ISL_526091\|2020- | ···················· | ···················· | ···················· | ···················· | ···················· | ···················· |
| G | 2155 hCoV-19/USA/WA-OHSU-0506/2020\|EPI_ISL_526092\|2020- | ···················· | ···················· | ···················· | ···················· | ···················· | ···················· |
| G | 2156 hCoV-19/USA/WA-OHSU-0512/2020\|EPI_ISL_526093\|2020- | ···················· | ···················· | ···················· | ···················· | ···················· | ···················· |
| G | 2157 hCoV-19/USA/WA-OHSU-0513/2020\|EPI_ISL_526094\|2020- | ···················· | ···················· | ···················· | ···················· | ···················· | ···················· |
| G | 2158 hCoV-19/USA/WA-OHSU-0515/2020\|EPI_ISL_526095\|2020- | ···················· | ···················· | ···················· | ···················· | ···················· | ···················· |
| G | 2159 hCoV-19/USA/WA-OHSU-0517/2020\|EPI_ISL_526096\|2020- | ···················· | ···················· | ···················· | ···················· | ···················· | ···················· |
| G | 2160 hCoV-19/USA/WA-OHSU-0520/2020\|EPI_ISL_526097\|2020- | ···················· | ···················· | ···················· | ···················· | ···················· | ···················· |
| G | 2161 hCoV-19/USA/WA-OHSU-0525/2020\|EPI_ISL_526098\|2020- | ···················· | ···················· | ···················· | ···················· | ···················· | ···················· |
| G | 2162 hCoV-19/USA/WA-OHSU-0821/2020\|EPI_ISL_526099\|2020- | ···················· | ···················· | ···················· | ···················· | ···················· | ···················· |
| G | 2163 hCoV-19/England/20162000304/2020\|EPI_ISL_527329\|20 | ···················· | ···················· | ···················· | ···················· | ···················· | ···················· |
| G | 2164 hCoV-19/England/20162002304/2020\|EPI_ISL_527331\|20 | ···················· | ···················· | ···················· | ···················· | ···················· | ···················· |
| G | 2165 hCoV-19/England/20162007704/2020\|EPI_ISL_527332\|20 | ···················· | ···················· | ···················· | ···················· | ···················· | ···················· |
| G | 2166 hCoV-19/England/20162007804/2020\|EPI_ISL_527333\|20 | ···················· | ···················· | ···················· | ···················· | ···················· | ···················· |
| G | 2167 hCoV-19/England/20162008304/2020\|EPI_ISL_527334\|20 | ···················· | ···················· | ···················· | ···················· | ···················· | ···················· |
| G | 2168 hCoV-19/England/20164002704/2020\|EPI_ISL_527335\|20 | ···················· | ···················· | ···················· | ···················· | ···················· | ···················· |
| G | 2169 hCoV-19/England/20166003004/2020\|EPI_ISL_527336\|20 | ···················· | ···················· | ···················· | ···················· | ···················· | ···················· |
| G | 2170 hCoV-19/England/20166003104/2020\|EPI_ISL_527337\|20 | ···················· | ···················· | ···················· | ···················· | ···················· | ···················· |
| G | 2172 hCoV-19/England/20179043204/2020\|EPI_ISL_527343\|20 | ···················· | ···················· | ···················· | ···················· | ···················· | ···················· |
| G | 2173 hCoV-19/England/20179043304/2020\|EPI_ISL_527344\|20 | ···················· | ···················· | ···················· | ···················· | ···················· | ···················· |
| G | 2174 hCoV-19/England/20179043504/2020\|EPI_ISL_527345\|20 | ···················· | ···················· | ···················· | ···················· | ···················· | ···················· |
| G | 2175 hCoV-19/England/20179043604/2020\|EPI_ISL_527346\|20 | ···················· | ···················· | ···················· | ···················· | ···················· | ···················· |
| G | 2176 hCoV-19/England/20179043704/2020\|EPI_ISL_527347\|20 | ···················· | ···················· | ···················· | ···················· | ···················· | ···················· |
| G | 2177 hCoV-19/England/20179043804/2020\|EPI_ISL_527348\|20 | ···················· | ···················· | ···················· | ···················· | ···················· | ···················· |
| G | 2178 hCoV-19/England/20179043904/2020\|EPI_ISL_527349\|20 | ···················· | ···················· | ···················· | ···················· | ···················· | ···················· |
| G | 2179 hCoV-19/England/20179044204/2020\|EPI_ISL_527351\|20 | ···················· | ···················· | ···················· | ···················· | ···················· | ···················· |
| G | 2180 hCoV-19/England/20179044304/2020\|EPI_ISL_527352\|20 | ···················· | ···················· | ···················· | ···················· | ···················· | ···················· |
| G | 2181 hCoV-19/England/20179044404/2020\|EPI_ISL_527353\|20 | ···················· | ···················· | ···················· | ···················· | ···················· | ···················· |
| G | 2182 hCoV-19/England/20179044604/2020\|EPI_ISL_527354\|20 | ···················· | ···················· | ···················· | ···················· | ···················· | ···················· |

| | | | | | | | |
|---|---|---|---|---|---|---|---|
| G | 2183 hCoV-19/England/20179044704/2020\|EPI_ISL_527355\|20 | .................... | .................... | .................... | .................... | .................... | .................... |
| G | 2184 hCoV-19/England/20179044804/2020\|EPI_ISL_527356\|20 | .................... | .................... | .................... | .................... | .................... | .................... |
| G | 2185 hCoV-19/England/20182001304/2020\|EPI_ISL_527357\|20 | .................... | .................... | .................... | .................... | .................... | .................... |
| G | 2186 hCoV-19/England/20186103704/2020\|EPI_ISL_527358\|20 | .................... | .................... | .................... | .................... | .................... | .................... |
| G | 2187 hCoV-19/Singapore/850/2020\|EPI_ISL_527359\|2020-08- | .................... | .................... | .................... | .................... | .................... | .................... |
| G | 2188 hCoV-19/Brazil/RJ-MD63-1a/2020\|EPI_ISL_528637\|2020 | .................... | .................... | .................... | .................... | .................... | .................... |
| G | 2189 hCoV-19/Brazil/RJ-MD63-1b/2020\|EPI_ISL_528638\|2020 | .................... | .................... | .................... | .................... | .................... | .................... |
| G | 2190 hCoV-19/USA/VA-DCLS-1180/2020\|EPI_ISL_528639\|2020- | .................... | .................... | .................... | .................... | .................... | .................... |
| G | 2191 hCoV-19/USA/VA-DCLS-1181/2020\|EPI_ISL_528640\|2020- | .................... | .................... | .................... | .................... | .................... | .................... |
| G | 2192 hCoV-19/USA/VA-DCLS-1182/2020\|EPI_ISL_528641\|2020- | .................... | .................... | .................... | .................... | .................... | .................... |
| G | 2193 hCoV-19/USA/VA-DCLS-1183/2020\|EPI_ISL_528642\|2020- | .................... | .................... | .................... | .................... | .................... | .................... |
| G | 2194 hCoV-19/USA/VA-DCLS-1184/2020\|EPI_ISL_528643\|2020- | .................... | .................... | .................... | .................... | .................... | .................... |
| G | 2195 hCoV-19/USA/VA-DCLS-1185/2020\|EPI_ISL_528644\|2020- | .................... | .................... | .................... | .................... | .................... | .................... |
| G | 2196 hCoV-19/USA/VA-DCLS-1186/2020\|EPI_ISL_528645\|2020- | .................... | .................... | .................... | .................... | .................... | .................... |
| G | 2197 hCoV-19/USA/VA-DCLS-1221/2020\|EPI_ISL_528646\|2020- | .................... | .A.................. | .................... | .................... | .................... | .................... |
| G | 2198 hCoV-19/USA/VA-DCLS-1222/2020\|EPI_ISL_528647\|2020- | .................... | .A.................. | .................... | .................... | .................... | .................... |
| G | 2199 hCoV-19/USA/VA-DCLS-1224/2020\|EPI_ISL_528648\|2020- | .................... | .................... | .................... | .................... | .................... | .................... |
| G | 2200 hCoV-19/USA/VA-DCLS-1225/2020\|EPI_ISL_528649\|2020- | .................... | .................... | .................... | .................... | .................... | .................... |
| G | 2201 hCoV-19/USA/VA-DCLS-1228/2020\|EPI_ISL_528650\|2020- | .................... | .A.................. | .................... | .................... | .................... | .................... |
| G | 2202 hCoV-19/USA/VA-DCLS-1232/2020\|EPI_ISL_528651\|2020- | .................... | .................... | .................... | .................... | .................... | .................... |
| G | 2203 hCoV-19/USA/VA-DCLS-1233/2020\|EPI_ISL_528652\|2020- | .................... | .................... | .................... | .................... | .................... | .................... |
| G | 2204 hCoV-19/USA/VA-DCLS-1235/2020\|EPI_ISL_528654\|2020- | .................... | .................... | .................... | .................... | .................... | .................... |
| G | 2205 hCoV-19/USA/VA-DCLS-1236/2020\|EPI_ISL_528655\|2020- | .................... | .................... | .................... | .................... | .................... | .................... |
| G | 2206 hCoV-19/USA/VA-DCLS-1237/2020\|EPI_ISL_528656\|2020- | .................... | .................... | .................... | .................... | .................... | .................... |
| G | 2207 hCoV-19/USA/VA-DCLS-1238/2020\|EPI_ISL_528657\|2020- | .................... | .................... | .................... | .................... | .................... | .................... |
| G | 2208 hCoV-19/USA/VA-DCLS-1239/2020\|EPI_ISL_528658\|2020- | .................... | .................... | .................... | .................... | .................... | .................... |
| G | 2209 hCoV-19/USA/VA-DCLS-1240/2020\|EPI_ISL_528659\|2020- | .................... | .................... | .................... | .................... | .................... | .................... |
| G | 2210 hCoV-19/USA/VA-DCLS-1241/2020\|EPI_ISL_528660\|2020- | .................... | .................... | .................... | .................... | .................... | .................... |
| G | 2211 hCoV-19/USA/VA-DCLS-1242/2020\|EPI_ISL_528661\|2020- | .................... | .................... | .................... | .................... | .................... | .................... |
| G | 2212 hCoV-19/USA/VA-DCLS-1243/2020\|EPI_ISL_528662\|2020- | .................... | .................... | .................... | .................... | .................... | .................... |
| G | 2213 hCoV-19/USA/VA-DCLS-1052/2020\|EPI_ISL_528663\|2020- | .................... | .................... | .................... | .................... | .................... | .................... |
| G | 2214 hCoV-19/USA/VA-DCLS-1056/2020\|EPI_ISL_528664\|2020- | .................... | .................... | .................... | .................... | .................... | .................... |
| G | 2215 hCoV-19/USA/VA-DCLS-1059/2020\|EPI_ISL_528665\|2020- | .................... | .................... | .................... | .................... | .................... | .................... |
| G | 2216 hCoV-19/USA/VA-DCLS-1060/2020\|EPI_ISL_528666\|2020- | .................... | .................... | .................... | .................... | .................... | .................... |
| G | 2217 hCoV-19/USA/VA-DCLS-1061/2020\|EPI_ISL_528667\|2020- | .................... | .................... | .................... | .................... | .................... | .................... |
| G | 2218 hCoV-19/USA/VA-DCLS-1064/2020\|EPI_ISL_528668\|2020- | .................... | ...A.A.............. | .................... | .................... | .................... | .................... |
| G | 2219 hCoV-19/USA/VA-DCLS-1067/2020\|EPI_ISL_528669\|2020- | .................... | .................... | .................... | .................... | .................... | .................... |
| G | 2220 hCoV-19/USA/VA-DCLS-1071/2020\|EPI_ISL_528670\|2020- | .................... | .................... | .................... | .................... | .................... | .................... |
| G | 2221 hCoV-19/USA/VA-DCLS-1072/2020\|EPI_ISL_528671\|2020- | .................... | .................... | .................... | .................... | .................... | .................... |
| G | 2222 hCoV-19/USA/VA-DCLS-1073/2020\|EPI_ISL_528672\|2020- | .................... | .................... | .................... | .................... | .................... | .................... |
| G | 2223 hCoV-19/USA/VA-DCLS-1074/2020\|EPI_ISL_528673\|2020- | .................... | .................... | .................... | .................... | .................... | .................... |
| G | 2224 hCoV-19/USA/VA-DCLS-1076/2020\|EPI_ISL_528674\|2020- | .................... | .................... | .................... | .................... | .................... | .................... |
| G | 2225 hCoV-19/USA/VA-DCLS-1077/2020\|EPI_ISL_528675\|2020- | .................... | .................... | .................... | .................... | .................... | .................... |
| G | 2226 hCoV-19/USA/VA-DCLS-1080/2020\|EPI_ISL_528676\|2020- | .................... | .................... | .................... | .................... | .................... | .................... |
| G | 2227 hCoV-19/USA/VA-DCLS-1081/2020\|EPI_ISL_528677\|2020- | .................... | .................... | .................... | .................... | .................... | .................... |
| G | 2228 hCoV-19/USA/VA-DCLS-1084/2020\|EPI_ISL_528678\|2020- | .................... | .................... | .................... | .................... | .................... | .................... |
| G | 2229 hCoV-19/USA/VA-DCLS-1087/2020\|EPI_ISL_528679\|2020- | .................... | .................... | .................... | .................... | .................... | .................... |
| G | 2230 hCoV-19/USA/VA-DCLS-1088/2020\|EPI_ISL_528680\|2020- | .......T............ | .................... | .................... | .................... | .................... | .................... |
| G | 2231 hCoV-19/USA/VA-DCLS-1089/2020\|EPI_ISL_528681\|2020- | .................... | .................... | .................... | .................... | .................... | .................... |
| G | 2232 hCoV-19/USA/VA-DCLS-1091/2020\|EPI_ISL_528682\|2020- | .................... | .................... | .................... | .................... | .................... | .................... |
| G | 2233 hCoV-19/India/DL-IGIB-77797/2020\|EPI_ISL_528683\|20 | .................... | .A.................. | .................... | .................... | .................... | ...A................ |
| G | 2234 hCoV-19/United Arab Emirates/12B/2020\|EPI_ISL_5286 | .................... | .................... | .................... | .................... | .................... | .................... |
| G | 2235 hCoV-19/United Arab Emirates/13/2020\|EPI_ISL_52868 | .................... | .................... | .................... | .................... | .................... | .................... |
| G | 2236 hCoV-19/United Arab Emirates/14B/2020\|EPI_ISL_5286 | .................... | .................... | .................... | .................... | .................... | .................... |
| G | 2237 hCoV-19/United Arab Emirates/16/2020\|EPI_ISL_52868 | .................... | .................... | .................... | .................... | .................... | .................... |
| G | 2238 hCoV-19/USA/VA-DCLS-1245/2020\|EPI_ISL_529938\|2020- | .................... | .................... | .................... | .................... | .................... | .................... |
| G | 2239 hCoV-19/USA/VA-DCLS-1246/2020\|EPI_ISL_529939\|2020- | .................... | .................... | .................... | .................... | .................... | .................... |
| G | 2240 hCoV-19/USA/VA-DCLS-1248/2020\|EPI_ISL_529940\|2020- | .................... | .................... | .................... | .................... | .................... | .................... |
| G | 2241 hCoV-19/USA/VA-DCLS-1249/2020\|EPI_ISL_529941\|2020- | .................... | .................... | .................... | .................... | .................... | .................... |
| G | 2242 hCoV-19/USA/VA-DCLS-1250/2020\|EPI_ISL_529942\|2020- | .................... | .................... | .................... | .................... | .................... | .................... |
| G | 2243 hCoV-19/USA/VA-DCLS-1252/2020\|EPI_ISL_529943\|2020- | .................... | .................... | .................... | .................... | .................... | .................... |
| G | 2244 hCoV-19/USA/VA-DCLS-1253/2020\|EPI_ISL_529944\|2020- | .................... | .................... | .................... | .................... | .................... | .................... |
| G | 2245 hCoV-19/USA/VA-DCLS-1254/2020\|EPI_ISL_529945\|2020- | .................... | .................... | .................... | .................... | .................... | .................... |
| G | 2246 hCoV-19/USA/VA-DCLS-1255/2020\|EPI_ISL_529946\|2020- | .................... | .................... | .................... | .................... | .................... | .................... |
| G | 2247 hCoV-19/USA/VA-DCLS-1257/2020\|EPI_ISL_529947\|2020- | .................... | .................... | .................... | .................... | .................... | .................... |
| G | 2248 hCoV-19/USA/VA-DCLS-1258/2020\|EPI_ISL_529948\|2020- | .................... | .................... | .................... | .................... | .................... | .................... |
| G | 2249 hCoV-19/USA/VA-DCLS-1259/2020\|EPI_ISL_529949\|2020- | .................... | .................... | .................... | .................... | .................... | .................... |
| G | 2250 hCoV-19/USA/VA-DCLS-1260/2020\|EPI_ISL_529950\|2020- | .................... | .................... | .................... | .................... | .................... | .................... |
| G | 2251 hCoV-19/USA/VA-DCLS-1261/2020\|EPI_ISL_529951\|2020- | .................... | .................... | .................... | .................... | .................... | .................... |
| G | 2252 hCoV-19/USA/VA-DCLS-1262/2020\|EPI_ISL_529952\|2020- | .................... | .................... | .................... | .................... | .................... | .................... |
| G | 2253 hCoV-19/USA/VA-DCLS-1263/2020\|EPI_ISL_529953\|2020- | .................... | .................... | .................... | .................... | .................... | .................... |
| G | 2254 hCoV-19/USA/VA-DCLS-1264/2020\|EPI_ISL_529954\|2020- | .................... | .................... | .................... | .................... | .................... | .................... |
| G | 2255 hCoV-19/USA/VA-DCLS-1265/2020\|EPI_ISL_529955\|2020- | .................... | .................... | .................... | .................... | .................... | .................... |
| G | 2256 hCoV-19/USA/VA-DCLS-1266/2020\|EPI_ISL_529956\|2020- | .................... | .................... | .................... | .................... | .................... | .................... |
| G | 2257 hCoV-19/USA/VA-DCLS-1268/2020\|EPI_ISL_529957\|2020- | .................... | .................... | .................... | .................... | .................... | .................... |
| G | 2258 hCoV-19/USA/VA-DCLS-1271/2020\|EPI_ISL_529958\|2020- | .................... | .................... | .................... | .................... | .................... | .................... |
| G | 2259 hCoV-19/USA/VA-DCLS-1273/2020\|EPI_ISL_529959\|2020- | .................... | .................... | .................... | .................... | .................... | .................... |
| G | 2260 hCoV-19/USA/VA-DCLS-1274/2020\|EPI_ISL_529960\|2020- | .................... | .................... | .................... | .................... | .................... | .................... |
| G | 2261 hCoV-19/Indonesia/JI-ITD-136N/2020\|EPI_ISL_529961\| | .................... | .................... | .................... | .................... | .................... | .................... |
| G | 2262 hCoV-19/Indonesia/JI-ITD-150Sp/2020\|EPI_ISL_529962 | .................... | .................... | .................... | .................... | .................... | .................... |

| | | | | | | | | |
|---|---|---|---|---|---|---|---|---|
| G | 2263 | hCoV-19/Indonesia/JI-ITD-17398NT/2020\|EPI_ISL_5299 | .................... | .................... | .................... | .................... | .................... | .................... |
| G | 2264 | hCoV-19/Indonesia/JI-ITD-1273V/2020\|EPI_ISL_529964 | .................... | .................... | .................... | .................... | .................... | .................... |
| G | 2265 | hCoV-19/Indonesia/JI-ITD-4859V/2020\|EPI_ISL_529965 | .................... | .................... | .................... | .................... | .................... | .................... |
| G | 2266 | hCoV-19/Indonesia/JI-ITD-7061V/2020\|EPI_ISL_529966 | .................... | .................... | .................... | .................... | .................... | .................... |
| G | 2267 | hCoV-19/Indonesia/JI-ITD-12202V/2020\|EPI_ISL_52996 | .................... | .................... | .................... | .................... | .................... | .................... |
| G | 2268 | hCoV-19/Spain/MD-H12-37/2020\|EPI_ISL_529972\|2020-0 | .................... | .................... | .................... | .................... | .................... | .................... |
| G | 2269 | hCoV-19/Spain/MD-H12-38/2020\|EPI_ISL_529973\|2020-0 | .................... | .................... | .................... | .................... | .................... | .................... |
| G | 2270 | hCoV-19/Spain/MD-H12-39/2020\|EPI_ISL_529974\|2020-0 | .................... | .................... | .................... | .................... | .................... | .................... |
| G | 2271 | hCoV-19/Spain/MD-H12-40/2020\|EPI_ISL_529975\|2020-0 | .................... | .................... | .................... | .................... | .................... | .................... |
| G | 2272 | hCoV-19/Spain/MD-H12-41/2020\|EPI_ISL_529976\|2020-0 | .................... | .................... | .................... | .................... | .................... | .................... |
| G | 2273 | hCoV-19/Spain/MD-H12-42/2020\|EPI_ISL_529977\|2020-0 | .................... | .................... | .................... | .................... | .................... | .................... |
| G | 2274 | hCoV-19/Spain/MD-H12-43/2020\|EPI_ISL_529978\|2020-0 | .................... | .................... | .................... | .................... | .................... | .................... |
| G | 2275 | hCoV-19/Spain/MD-H12-46/2020\|EPI_ISL_529979\|2020-0 | .................... | .................... | .................... | .................... | .................... | .................... |
| G | 2276 | hCoV-19/Spain/MD-H12-47/2020\|EPI_ISL_529980\|2020-0 | .................... | .................... | .................... | .................... | .................... | .................... |
| G | 2277 | hCoV-19/Spain/MD-H12-48/2020\|EPI_ISL_529981\|2020-0 | .................... | .................... | .................... | .................... | .................... | .................... |
| G | 2278 | hCoV-19/Spain/MD-H12-49/2020\|EPI_ISL_529982\|2020-0 | .................... | .................... | .................... | .................... | .................... | .................... |
| G | 2279 | hCoV-19/Spain/MD-H12-50/2020\|EPI_ISL_529983\|2020-0 | .................... | .................... | .................... | .................... | .................... | .................... |
| G | 2280 | hCoV-19/Spain/MD-H12-52/2020\|EPI_ISL_529984\|2020-0 | .................... | .................... | .................... | .................... | .................... | .................... |
| G | 2281 | hCoV-19/Spain/MD-H12-53/2020\|EPI_ISL_529985\|2020-0 | .................... | .................... | .................... | .................... | .................... | .................... |
| G | 2282 | hCoV-19/Spain/MD-H12-54/2020\|EPI_ISL_529986\|2020-0 | .................... | .................... | .................... | .................... | .................... | .................... |
| G | 2283 | hCoV-19/Spain/MD-H12-55/2020\|EPI_ISL_529987\|2020-0 | .................... | .................... | .................... | .................... | .................... | .................... |
| G | 2284 | hCoV-19/Spain/MD-H12-56/2020\|EPI_ISL_529988\|2020-0 | .................... | .................... | .................... | .................... | .................... | .................... |
| G | 2285 | hCoV-19/Spain/MD-H12-57/2020\|EPI_ISL_529989\|2020-0 | .................... | .................... | .................... | .................... | .................... | .................... |
| G | 2286 | hCoV-19/Spain/MD-H12-58/2020\|EPI_ISL_529990\|2020-0 | .................... | .................... | .................... | .................... | .................... | .................... |
| G | 2287 | hCoV-19/Spain/MD-H12-59/2020\|EPI_ISL_529991\|2020-0 | .................... | .A.................. | .................... | .................... | .................... | .................... |
| G | 2288 | hCoV-19/Spain/MD-H12-61/2020\|EPI_ISL_529992\|2020-0 | .................... | .................... | .................... | .................... | .................... | .................... |
| G | 2289 | hCoV-19/Spain/MD-H12-62/2020\|EPI_ISL_529993\|2020-0 | .................... | .................... | .................... | .................... | .................... | .................... |
| G | 2290 | hCoV-19/Spain/MD-H12-63/2020\|EPI_ISL_529994\|2020-0 | .................... | .................... | .................... | .................... | .................... | .................... |
| G | 2291 | hCoV-19/Spain/MD-H12-64/2020\|EPI_ISL_529995\|2020-0 | .................... | .................... | .................... | .................... | .................... | .................... |
| G | 2292 | hCoV-19/Spain/MD-H12-65/2020\|EPI_ISL_529996\|2020-0 | .................... | .................... | .................... | .................... | .................... | .................... |
| G | 2293 | hCoV-19/Spain/MD-H12-66/2020\|EPI_ISL_529997\|2020-0 | .................... | .................... | .................... | .................... | .................... | .................... |
| G | 2294 | hCoV-19/Spain/MD-H12-67/2020\|EPI_ISL_529998\|2020-0 | .................... | .................... | .................... | .................... | .................... | .................... |
| G | 2295 | hCoV-19/Spain/MD-H12-68/2020\|EPI_ISL_529999\|2020-0 | .................... | .................... | .................... | .................... | .................... | .................... |
| G | 2296 | hCoV-19/Japan/TK-YE63319/2020\|EPI_ISL_582135\|2020- | .................... | .................... | .................... | .................... | .................... | .................... |
| G | 2297 | hCoV-19/Japan/TK-YE63557/2020\|EPI_ISL_582136\|2020- | .................... | .................... | .................... | .................... | .................... | .................... |
| G | 2298 | hCoV-19/Japan/TK-YE63557d/2020\|EPI_ISL_582137\|2020 | .................... | .................... | .................... | .................... | .................... | .................... |
| G | 2299 | hCoV-19/Japan/TK-YE63558/2020\|EPI_ISL_582138\|2020- | .................... | .................... | .................... | .................... | .................... | .................... |
| G | 2300 | hCoV-19/Japan/TK-YE63558d/2020\|EPI_ISL_582139\|2020 | .................... | .................... | .................... | .................... | .................... | .................... |
| G | 2301 | hCoV-19/Japan/TK-YE63736/2020\|EPI_ISL_582140\|2020- | .................... | .................... | .................... | .................... | .................... | .................... |
| G | 2302 | hCoV-19/Japan/TK-YE63736d/2020\|EPI_ISL_582141\|2020 | .................... | .................... | .................... | .................... | .................... | .................... |
| G | 2303 | hCoV-19/Japan/TK-YE63738/2020\|EPI_ISL_582142\|2020- | .................... | .................... | .................... | .................... | .................... | .................... |
| G | 2304 | hCoV-19/Japan/TK-YE63738d/2020\|EPI_ISL_582143\|2020 | .................... | .................... | .................... | .................... | .................... | .................... |
| G | 2305 | hCoV-19/Australia/VIC6259/2020\|EPI_ISL_518250\|2020 | .................... | .................... | .................... | .................... | .................... | .................... |
| G | 2306 | hCoV-19/Australia/VIC6260/2020\|EPI_ISL_518251\|2020 | .................... | .................... | .................... | .................... | .................... | .................... |
| G | 2307 | hCoV-19/Australia/VIC6261/2020\|EPI_ISL_518252\|2020 | .................... | .................... | .................... | .................... | .................... | .................... |
| G | 2308 | hCoV-19/Australia/VIC6262/2020\|EPI_ISL_518253\|2020 | .................... | .................... | .................... | .................... | .................... | .................... |
| G | 2309 | hCoV-19/Australia/VIC6263/2020\|EPI_ISL_518254\|2020 | .................... | .................... | .................... | .................... | .................... | .................... |
| G | 2310 | hCoV-19/Australia/VIC6265/2020\|EPI_ISL_518255\|2020 | .................... | .................... | .................... | .................... | .................... | .................... |
| G | 2311 | hCoV-19/Australia/VIC6266/2020\|EPI_ISL_518256\|2020 | .................... | .................... | .................... | .................... | .................... | .................... |
| G | 2312 | hCoV-19/Australia/VIC6267/2020\|EPI_ISL_518257\|2020 | .................... | .................... | .................... | .................... | .................... | .................... |
| G | 2313 | hCoV-19/Australia/VIC6268/2020\|EPI_ISL_518258\|2020 | .................... | .................... | .................... | .................... | .................... | .................... |
| G | 2314 | hCoV-19/Australia/VIC6269/2020\|EPI_ISL_518259\|2020 | .................... | .................... | .................... | .................... | .................... | .................... |
| G | 2315 | hCoV-19/Australia/VIC6270/2020\|EPI_ISL_518260\|2020 | .................... | .................... | .................... | .................... | .................... | .................... |
| G | 2316 | hCoV-19/Australia/VIC6271/2020\|EPI_ISL_518261\|2020 | .................... | .................... | .................... | .................... | .................... | .................... |
| G | 2317 | hCoV-19/Australia/VIC6272/2020\|EPI_ISL_518262\|2020 | .................... | .................... | .................... | .................... | .................... | .................... |
| G | 2318 | hCoV-19/Australia/VIC6273/2020\|EPI_ISL_518263\|2020 | .................... | .................... | .................... | .................... | .................... | .................... |
| G | 2319 | hCoV-19/Australia/VIC6275/2020\|EPI_ISL_518264\|2020 | .................... | .................... | .................... | .................... | .................... | .................... |
| G | 2320 | hCoV-19/Australia/VIC6276/2020\|EPI_ISL_518265\|2020 | .................... | .................... | .................... | .................... | .................... | .................... |
| G | 2321 | hCoV-19/Australia/VIC6277/2020\|EPI_ISL_518266\|2020 | .................... | .................... | .................... | .................... | .................... | .................... |
| G | 2322 | hCoV-19/Australia/VIC6278/2020\|EPI_ISL_518267\|2020 | .................... | .................... | .................... | .................... | .................... | .................... |
| G | 2323 | hCoV-19/Australia/VIC6279/2020\|EPI_ISL_518268\|2020 | .................... | .................... | .................... | .................... | .................... | .................... |
| G | 2324 | hCoV-19/Australia/VIC6281/2020\|EPI_ISL_518269\|2020 | .................... | .................... | .................... | .................... | .................... | .................... |
| G | 2325 | hCoV-19/Australia/VIC6282/2020\|EPI_ISL_518270\|2020 | .................... | .................... | .................... | .................... | .................... | .................... |
| G | 2326 | hCoV-19/Australia/VIC6283/2020\|EPI_ISL_518271\|2020 | .................... | .................... | .................... | .................... | .................... | .................... |
| G | 2327 | hCoV-19/Australia/VIC6284/2020\|EPI_ISL_518272\|2020 | .................... | .................... | .................... | .................... | .................... | .................... |
| G | 2328 | hCoV-19/Australia/VIC6285/2020\|EPI_ISL_518273\|2020 | .................... | .................... | .................... | .................... | .................... | .................... |
| G | 2329 | hCoV-19/Australia/VIC6286/2020\|EPI_ISL_518274\|2020 | .................... | .................... | .................... | .................... | .................... | .................... |
| G | 2330 | hCoV-19/Australia/VIC6287/2020\|EPI_ISL_518275\|2020 | .................... | .................... | .................... | .................... | .................... | .................... |
| G | 2331 | hCoV-19/Australia/VIC6288/2020\|EPI_ISL_518276\|2020 | .................... | .................... | .................... | .................... | .................... | .................... |
| G | 2332 | hCoV-19/Australia/VIC6289/2020\|EPI_ISL_518277\|2020 | .................... | .................... | .................... | .................... | .................... | .................... |
| G | 2333 | hCoV-19/Australia/VIC6290/2020\|EPI_ISL_518278\|2020 | .................... | .................... | .................... | .................... | .................... | .................... |
| G | 2334 | hCoV-19/Australia/VIC6294/2020\|EPI_ISL_518282\|2020 | .................... | .................... | .................... | .................... | .................... | .................... |
| G | 2335 | hCoV-19/Australia/VIC6295/2020\|EPI_ISL_518283\|2020 | .................... | .................... | .................... | .................... | .................... | .................... |
| G | 2336 | hCoV-19/Australia/VIC6296/2020\|EPI_ISL_518284\|2020 | .................... | .................... | .................... | .................... | .................... | .................... |
| G | 2337 | hCoV-19/Australia/VIC6297/2020\|EPI_ISL_518285\|2020 | .................... | .................... | .................... | .................... | .................... | .................... |
| G | 2338 | hCoV-19/Australia/VIC6299/2020\|EPI_ISL_518287\|2020 | .................... | .................... | .................... | .................... | .................... | .................... |
| G | 2339 | hCoV-19/Australia/VIC6300/2020\|EPI_ISL_518288\|2020 | .................... | .................... | .................... | .................... | .................... | .................... |
| G | 2340 | hCoV-19/Australia/VIC6301/2020\|EPI_ISL_518289\|2020 | .................... | .................... | .................... | .................... | .................... | .................... |
| G | 2341 | hCoV-19/Australia/VIC6302/2020\|EPI_ISL_518290\|2020 | .................... | .................... | .................... | .................... | .................... | .................... |
| G | 2342 | hCoV-19/Australia/VIC6304/2020\|EPI_ISL_518292\|2020 | .................... | .................... | .................... | .................... | .................... | .................... |

| | | | | | | | | |
|---|---|---|---|---|---|---|---|---|
| G | 2343 | hCoV-19/Australia/VIC6305/2020\|EPI_ISL_518293\|2020 | ···················· | ···················· | ····················· | ···················· | ···················· | ···················· |
| G | 2344 | hCoV-19/Australia/VIC6306/2020\|EPI_ISL_518294\|2020 | ···················· | ···················· | ····················· | ···················· | ···················· | ···················· |
| G | 2345 | hCoV-19/Australia/VIC6307/2020\|EPI_ISL_518295\|2020 | ···················· | ···················· | ····················· | ···················· | ···················· | ···················· |
| G | 2346 | hCoV-19/Australia/VIC6308/2020\|EPI_ISL_518296\|2020 | ···················· | ···················· | ····················· | ···················· | ···················· | ···················· |
| G | 2347 | hCoV-19/Australia/VIC6309/2020\|EPI_ISL_518297\|2020 | ···················· | ···················· | ····················· | ···················· | ···················· | ···················· |
| G | 2348 | hCoV-19/Australia/VIC6310/2020\|EPI_ISL_518298\|2020 | ···················· | ···················· | ····················· | ···················· | ···················· | ···················· |
| G | 2349 | hCoV-19/Australia/VIC6311/2020\|EPI_ISL_518299\|2020 | ···················· | ···················· | ····················· | ···················· | ···················· | ···················· |
| G | 2350 | hCoV-19/Australia/VIC4252/2020\|EPI_ISL_519580\|2020 | ···················· | ···················· | ····················· | ···················· | ···················· | ···················· |
| G | 2351 | hCoV-19/Australia/VIC4253/2020\|EPI_ISL_519581\|2020 | ···················· | ···················· | ····················· | ···················· | ···················· | ···················· |
| G | 2352 | hCoV-19/Australia/VIC4254/2020\|EPI_ISL_519582\|2020 | ···················· | ···················· | ····················· | ···················· | ···················· | ···················· |
| G | 2353 | hCoV-19/Australia/VIC4255/2020\|EPI_ISL_519583\|2020 | ···················· | ···················· | ····················· | ···················· | ···················· | ···················· |
| G | 2354 | hCoV-19/Australia/VIC4256/2020\|EPI_ISL_519584\|2020 | ···················· | ···················· | ····················· | ···················· | ···················· | ···················· |
| G | 2355 | hCoV-19/Australia/VIC4257/2020\|EPI_ISL_519585\|2020 | ···················· | ···················· | ····················· | ···················· | ···················· | ···················· |
| G | 2356 | hCoV-19/Australia/VIC4258/2020\|EPI_ISL_519586\|2020 | ···················· | ···················· | ····················· | ···················· | ···················· | ···················· |
| G | 2357 | hCoV-19/Australia/VIC4259/2020\|EPI_ISL_519587\|2020 | ···················· | ···················· | ····················· | ···················· | ···················· | ···················· |
| G | 2358 | hCoV-19/Australia/VIC4260/2020\|EPI_ISL_519588\|2020 | ···················· | ···················· | ····················· | ···················· | ···················· | ···················· |
| G | 2359 | hCoV-19/Australia/VIC4261/2020\|EPI_ISL_519589\|2020 | ···················· | ···················· | ····················· | ···················· | ···················· | ···················· |
| G | 2360 | hCoV-19/Australia/VIC4262/2020\|EPI_ISL_519590\|2020 | ···················· | ···················· | ····················· | ···················· | ···················· | ···················· |
| G | 2361 | hCoV-19/Australia/VIC4263/2020\|EPI_ISL_519591\|2020 | ···················· | ···················· | ····················· | ···················· | ···················· | ···················· |
| G | 2362 | hCoV-19/Australia/VIC4264/2020\|EPI_ISL_519592\|2020 | ···················· | ···················· | ····················· | ···················· | ···················· | ···················· |
| G | 2363 | hCoV-19/Australia/VIC4265/2020\|EPI_ISL_519593\|2020 | ···················· | ···················· | ····················· | ···················· | ···················· | ···················· |
| G | 2364 | hCoV-19/Australia/VIC4266/2020\|EPI_ISL_519594\|2020 | ···················· | ···················· | ····················· | ···················· | ···················· | ···················· |
| G | 2365 | hCoV-19/Australia/VIC4267/2020\|EPI_ISL_519595\|2020 | ···················· | ···················· | ····················· | ···················· | ···················· | ···················· |
| G | 2366 | hCoV-19/Australia/VIC4268/2020\|EPI_ISL_519596\|2020 | ···················· | ···················· | ····················· | ···················· | ···················· | ···················· |
| G | 2367 | hCoV-19/Australia/VIC4269/2020\|EPI_ISL_519597\|2020 | ···················· | ···················· | ····················· | ···················· | ···················· | ···················· |
| G | 2368 | hCoV-19/Australia/VIC4270/2020\|EPI_ISL_519598\|2020 | ···················· | ···················· | ····················· | ···················· | ···················· | ···················· |
| G | 2369 | hCoV-19/Australia/VIC4271/2020\|EPI_ISL_519599\|2020 | ···················· | ···················· | ····················· | ···················· | ···················· | ···················· |
| G | 2370 | hCoV-19/Australia/VIC3853/2020\|EPI_ISL_520566\|2020 | ···················· | ···················· | ····················· | ···················· | ···················· | ···················· |
| G | 2371 | hCoV-19/Australia/VIC3854/2020\|EPI_ISL_520567\|2020 | ···················· | ···················· | ····················· | ···················· | ···················· | ···················· |
| G | 2372 | hCoV-19/Australia/VIC3855/2020\|EPI_ISL_520568\|2020 | ···················· | ···················· | ····················· | ···················· | ···················· | ···················· |
| G | 2373 | hCoV-19/Australia/VIC3856/2020\|EPI_ISL_520569\|2020 | ···················· | ···················· | ····················· | ···················· | ···················· | ···················· |
| G | 2374 | hCoV-19/Australia/VIC3857/2020\|EPI_ISL_520570\|2020 | ···················· | ···················· | ····················· | ···················· | ···················· | ···················· |
| G | 2375 | hCoV-19/Australia/VIC3858/2020\|EPI_ISL_520571\|2020 | ···················· | ···················· | ····················· | ···················· | ···················· | ···················· |
| G | 2376 | hCoV-19/Australia/VIC3859/2020\|EPI_ISL_520572\|2020 | ···················· | ···················· | ····················· | ···················· | ···················· | ···················· |
| G | 2377 | hCoV-19/Australia/VIC3860/2020\|EPI_ISL_520573\|2020 | ···················· | ···················· | ····················· | ···················· | ···················· | ···················· |
| G | 2378 | hCoV-19/Australia/VIC3861/2020\|EPI_ISL_520574\|2020 | ···················· | ···················· | ····················· | ···················· | ···················· | ···················· |
| G | 2379 | hCoV-19/Australia/VIC3862/2020\|EPI_ISL_520575\|2020 | ···················· | ···················· | ····················· | ···················· | ···················· | ···················· |
| G | 2380 | hCoV-19/Australia/VIC3863/2020\|EPI_ISL_520576\|2020 | ···················· | ···················· | ····················· | ···················· | ···················· | ···················· |
| G | 2381 | hCoV-19/Australia/VIC3864/2020\|EPI_ISL_520577\|2020 | ···················· | ···················· | ····················· | ···················· | ···················· | ···················· |
| G | 2382 | hCoV-19/Australia/VIC3865/2020\|EPI_ISL_520578\|2020 | ···················· | ···················· | ····················· | ···················· | ···················· | ···················· |
| G | 2383 | hCoV-19/Australia/VIC3866/2020\|EPI_ISL_520579\|2020 | ···················· | ···················· | ····················· | ···················· | ···················· | ···················· |
| G | 2384 | hCoV-19/Australia/VIC3867/2020\|EPI_ISL_520580\|2020 | ···················· | ···················· | ····················· | ···················· | ···················· | ···················· |
| G | 2385 | hCoV-19/Australia/VIC3868/2020\|EPI_ISL_520581\|2020 | ···················· | ···················· | ····················· | ···················· | ···················· | ···················· |
| G | 2386 | hCoV-19/Australia/VIC3869/2020\|EPI_ISL_520582\|2020 | ···················· | ···················· | ····················· | ···················· | ···················· | ···················· |
| G | 2387 | hCoV-19/Australia/VIC3870/2020\|EPI_ISL_520583\|2020 | ···················· | ···················· | ····················· | ···················· | ···················· | ···················· |
| G | 2388 | hCoV-19/Australia/VIC3871/2020\|EPI_ISL_520584\|2020 | ···················· | ···················· | ····················· | ···················· | ···················· | ···················· |
| G | 2389 | hCoV-19/Australia/VIC3872/2020\|EPI_ISL_520585\|2020 | ···················· | ···················· | ····················· | ···················· | ···················· | ···················· |
| G | 2390 | hCoV-19/Australia/VIC3873/2020\|EPI_ISL_520586\|2020 | ···················· | ···················· | ····················· | ···················· | ···················· | ···················· |
| G | 2391 | hCoV-19/Australia/VIC3874/2020\|EPI_ISL_520587\|2020 | ···················· | ···················· | ····················· | ···················· | ···················· | ···················· |
| G | 2392 | hCoV-19/Australia/VIC3875/2020\|EPI_ISL_520588\|2020 | ···················· | ···················· | ····················· | ···················· | ···················· | ···················· |
| G | 2393 | hCoV-19/Australia/VIC3876/2020\|EPI_ISL_520589\|2020 | ···················· | ···················· | ····················· | ···················· | ···················· | ···················· |
| G | 2394 | hCoV-19/Australia/VIC3877/2020\|EPI_ISL_520590\|2020 | ···················· | ···················· | ····················· | ···················· | ···················· | ···················· |
| G | 2395 | hCoV-19/Australia/VIC3878/2020\|EPI_ISL_520591\|2020 | ···················· | ···················· | ····················· | ···················· | ···················· | ···················· |
| G | 2396 | hCoV-19/Australia/VIC3879/2020\|EPI_ISL_520592\|2020 | ···················· | ···················· | ····················· | ···················· | ···················· | ···················· |
| G | 2397 | hCoV-19/Australia/VIC3880/2020\|EPI_ISL_520593\|2020 | ···················· | ···················· | ····················· | ···················· | ···················· | ···················· |
| G | 2398 | hCoV-19/Australia/VIC3881/2020\|EPI_ISL_520594\|2020 | ···················· | ···················· | ····················· | ···················· | ···················· | ···················· |
| G | 2399 | hCoV-19/Australia/VIC3883/2020\|EPI_ISL_520596\|2020 | ···················· | ···················· | ····················· | ···················· | ···················· | ···················· |
| G | 2400 | hCoV-19/Australia/VIC3884/2020\|EPI_ISL_520597\|2020 | ···················· | ···················· | ····················· | ···················· | ···················· | ···················· |
| G | 2401 | hCoV-19/Australia/VIC3885/2020\|EPI_ISL_520598\|2020 | ···················· | ···················· | ····················· | ···················· | ···················· | ···················· |
| G | 2402 | hCoV-19/Australia/VIC3886/2020\|EPI_ISL_520599\|2020 | ···················· | ···················· | ····················· | ···················· | ···················· | ···················· |
| G | 2405 | hCoV-19/Netherlands/ZH-EMC-464/2020\|EPI_ISL_523800 | ···················· | ···················· | ····················· | ···················· | ···················· | ···················· |
| G | 2406 | hCoV-19/Netherlands/ZH-EMC-466/2020\|EPI_ISL_523802 | ···················· | ···················· | ····················· | ···················· | ···················· | ···················· |
| G | 2407 | hCoV-19/Netherlands/ZH-EMC-467/2020\|EPI_ISL_523803 | ···················· | ···················· | ····················· | ···················· | ···················· | ···················· |
| G | 2408 | hCoV-19/Netherlands/ZH-EMC-468/2020\|EPI_ISL_523804 | ···················· | ···················· | ····················· | ···················· | ···················· | ···················· |
| G | 2409 | hCoV-19/Netherlands/ZH-EMC-469/2020\|EPI_ISL_523805 | ···················· | ···················· | ····················· | ···················· | ···················· | ···················· |
| G | 2410 | hCoV-19/Netherlands/ZH-EMC-470/2020\|EPI_ISL_523806 | ···················· | ·A·················· | ····················· | ···················· | ···················· | ···················· |
| G | 2411 | hCoV-19/Netherlands/ZH-EMC-471/2020\|EPI_ISL_523807 | ···················· | ···················· | ····················· | ···················· | ···················· | ···················· |
| G | 2412 | hCoV-19/Netherlands/ZH-EMC-472/2020\|EPI_ISL_523808 | ···················· | ···················· | ····················· | ···················· | ···················· | ···················· |
| G | 2413 | hCoV-19/Netherlands/ZH-EMC-473/2020\|EPI_ISL_523809 | ···················· | ···················· | ····················· | ···················· | ···················· | ···················· |
| G | 2414 | hCoV-19/Peru/LAL-INS-070/2020\|EPI_ISL_523810\|2020- | ···················· | ···················· | ····················· | ···················· | ···················· | ···················· |
| G | 2415 | hCoV-19/USA/UT-08308/2020\|EPI_ISL_524104\|2020-05-2 | ···················· | ···················· | ····················· | ···················· | ···················· | ···················· |
| G | 2416 | hCoV-19/USA/UT-08312/2020\|EPI_ISL_524108\|2020-05-1 | ···················· | ···················· | ····················· | ···················· | ···················· | ···················· |
| G | 2417 | hCoV-19/USA/UT-08313/2020\|EPI_ISL_524109\|2020-05-1 | ···················· | ···················· | ····················· | ···················· | ···················· | ···················· |
| G | 2418 | hCoV-19/USA/UT-08316/2020\|EPI_ISL_524111\|2020-05-1 | ···················· | ···················· | ····················· | ···················· | ···················· | ···················· |
| G | 2419 | hCoV-19/USA/UT-08318/2020\|EPI_ISL_524113\|2020-05-1 | ···················· | ···················· | ····················· | ···················· | ···················· | ···················· |
| G | 2420 | hCoV-19/USA/UT-08319/2020\|EPI_ISL_524114\|2020-05-1 | ···················· | ···················· | ····················· | ···················· | ···················· | ···················· |
| G | 2421 | hCoV-19/USA/UT-08320/2020\|EPI_ISL_524115\|2020-05-1 | ···················· | ···················· | ····················· | ···················· | ···················· | ···················· |
| G | 2422 | hCoV-19/USA/UT-08324/2020\|EPI_ISL_524117\|2020-05-1 | ···················· | ···················· | ····················· | ···················· | ···················· | ···················· |
| G | 2423 | hCoV-19/USA/UT-08325/2020\|EPI_ISL_524118\|2020-05-1 | ···················· | ···················· | ····················· | ···················· | ···················· | ···················· |
| G | 2424 | hCoV-19/USA/UT-08330/2020\|EPI_ISL_524121\|2020-05-1 | ···················· | ···················· | ····················· | ···················· | ···················· | ···················· |

| | | | | | | | |
|---|---|---|---|---|---|---|---|
| G | 2425 hCoV-19/USA/UT-08331/2020\|EPI_ISL_524122\|2020-05-1 | ···················· | ···················· | ···················· | ···················· | ···················· | ···················· |
| G | 2426 hCoV-19/USA/UT-08332/2020\|EPI_ISL_524123\|2020-05-2 | ···················· | ···················· | ···················· | ···················· | ···················· | ···················· |
| G | 2427 hCoV-19/USA/UT-08333/2020\|EPI_ISL_524124\|2020-05-2 | ···················· | ···················· | ···················· | ···················· | ···················· | ···················· |
| G | 2428 hCoV-19/USA/UT-08336/2020\|EPI_ISL_524125\|2020-05-1 | ···················· | ···················· | ···················· | ···················· | ···················· | ···················· |
| G | 2429 hCoV-19/USA/UT-08337/2020\|EPI_ISL_524126\|2020-05-1 | ···················· | ···················· | ···················· | ···················· | ···················· | ···················· |
| G | 2430 hCoV-19/USA/UT-08339/2020\|EPI_ISL_524127\|2020-05-1 | ···················· | ···················· | ···················· | ···················· | ···················· | ···················· |
| G | 2431 hCoV-19/USA/UT-08340/2020\|EPI_ISL_524128\|2020-05-1 | ···················· | ···················· | ···················· | ···················· | ···················· | ···················· |
| G | 2432 hCoV-19/USA/UT-08341/2020\|EPI_ISL_524129\|2020-05-1 | ···················· | ···················· | ···················· | ···················· | ···················· | ···················· |
| G | 2433 hCoV-19/Ecuador/USFQ-163/2020\|EPI_ISL_525430\|2020- | ···················· | ···················· | ···················· | ···················· | ···················· | ···················· |
| G | 2434 hCoV-19/Ecuador/USFQ-184/2020\|EPI_ISL_525435\|2020- | ···················· | ···················· | ···················· | ···················· | ···················· | ···················· |
| G | 2435 hCoV-19/Ecuador/USFQ-186/2020\|EPI_ISL_525436\|2020- | ···················· | ···················· | ···················· | ···················· | ···················· | ···················· |
| G | 2436 hCoV-19/USA/OR-OHSU-1801/2020\|EPI_ISL_526030\|2020- | ···················· | ·A·················· | ···················· | ···················· | ···················· | ···················· |
| G | 2437 hCoV-19/USA/OR-OHSU-1803/2020\|EPI_ISL_526031\|2020- | ···················· | ·A·················· | ···················· | ···················· | ···················· | ···················· |
| G | 2438 hCoV-19/USA/OR-OHSU-1807/2020\|EPI_ISL_526032\|2020- | ···················· | ·A·················· | ···················· | ···················· | ···················· | ···················· |
| G | 2439 hCoV-19/USA/OR-OHSU-1809/2020\|EPI_ISL_526033\|2020- | ···················· | ·A·················· | ···················· | ···················· | ···················· | ···················· |
| G | 2440 hCoV-19/USA/OR-OHSU-1811/2020\|EPI_ISL_526034\|2020- | ···················· | ···················· | ···················· | ···················· | ···················· | ···················· |
| G | 2441 hCoV-19/USA/OR-OHSU-1813/2020\|EPI_ISL_526035\|2020- | ···················· | ·A·················· | ···················· | ···················· | ···················· | ···················· |
| G | 2442 hCoV-19/USA/OR-OHSU-1815/2020\|EPI_ISL_526036\|2020- | ···················· | ···················· | ···················· | ···················· | ···················· | ···················· |
| G | 2443 hCoV-19/USA/OR-OHSU-1817/2020\|EPI_ISL_526037\|2020- | ···················· | ·A·················· | ···················· | ···················· | ···················· | ···················· |
| G | 2444 hCoV-19/USA/OR-OHSU-1823/2020\|EPI_ISL_526038\|2020- | ···················· | ···················· | ···················· | ···················· | ···················· | ···················· |
| G | 2445 hCoV-19/USA/OR-OHSU-1825/2020\|EPI_ISL_526039\|2020- | ···················· | ···················· | ···················· | ···················· | ···················· | ···················· |
| G | 2446 hCoV-19/USA/OR-OHSU-1827/2020\|EPI_ISL_526040\|2020- | ···················· | ···················· | ···················· | ···················· | ···················· | ···················· |
| G | 2447 hCoV-19/USA/OR-OHSU-1833/2020\|EPI_ISL_526041\|2020- | ···················· | ···················· | ···················· | ···················· | ···················· | ···················· |
| G | 2448 hCoV-19/USA/OR-OHSU-1837/2020\|EPI_ISL_526043\|2020- | ···················· | ···················· | ···················· | ···················· | ···················· | ···················· |
| G | 2449 hCoV-19/USA/OR-OHSU-1839/2020\|EPI_ISL_526044\|2020- | ···················· | ···················· | ···················· | ···················· | ···················· | ···················· |
| G | 2450 hCoV-19/USA/OR-OHSU-1843/2020\|EPI_ISL_526045\|2020- | ···················· | ·A·················· | ···················· | ···················· | ···················· | ···················· |
| G | 2451 hCoV-19/USA/OR-OHSU-1847/2020\|EPI_ISL_526046\|2020- | ···················· | ···················· | ···················· | ···················· | ···················· | ···················· |
| G | 2452 hCoV-19/USA/OR-OHSU-1849/2020\|EPI_ISL_526047\|2020- | ···················· | ···················· | ···················· | ···················· | ···················· | ···················· |
| G | 2453 hCoV-19/USA/OR-OHSU-1855/2020\|EPI_ISL_526048\|2020- | ···················· | ···················· | ···················· | ···················· | ···················· | ···················· |
| G | 2454 hCoV-19/USA/OR-OHSU-1857/2020\|EPI_ISL_526049\|2020- | ···················· | ···················· | ···················· | ···················· | ···················· | ···················· |
| G | 2455 hCoV-19/USA/OR-OHSU-1859/2020\|EPI_ISL_526050\|2020- | ···················· | ···················· | ···················· | ···················· | ···················· | ···················· |
| G | 2456 hCoV-19/USA/OR-OHSU-1863/2020\|EPI_ISL_526051\|2020- | ···················· | ···················· | ···················· | ···················· | ···················· | ···················· |
| G | 2457 hCoV-19/USA/OR-OHSU-1865/2020\|EPI_ISL_526052\|2020- | ···················· | ·A·················· | ···················· | ···················· | ···················· | ···················· |
| G | 2458 hCoV-19/USA/OR-OHSU-1867/2020\|EPI_ISL_526053\|2020- | ···················· | ·A·················· | ···················· | ···················· | ···················· | ···················· |
| G | 2459 hCoV-19/USA/OR-OHSU-1873/2020\|EPI_ISL_526054\|2020- | ···················· | ···················· | ···················· | ···················· | ···················· | ···················· |
| G | 2460 hCoV-19/USA/OR-OHSU-1875/2020\|EPI_ISL_526055\|2020- | ···················· | ···················· | ···················· | ···················· | ···················· | ···················· |
| G | 2461 hCoV-19/USA/OR-OHSU-1877/2020\|EPI_ISL_526056\|2020- | ···················· | ···················· | ···················· | ···················· | ···················· | ···················· |
| G | 2462 hCoV-19/USA/OR-OHSU-1879/2020\|EPI_ISL_526057\|2020- | ···················· | ···················· | ···················· | ···················· | ···················· | ···················· |
| G | 2463 hCoV-19/USA/OR-OHSU-1881/2020\|EPI_ISL_526058\|2020- | ···················· | ···················· | ···················· | ···················· | ···················· | ···················· |
| G | 2464 hCoV-19/USA/OR-OHSU-1883/2020\|EPI_ISL_526059\|2020- | ···················· | ·A·················· | ···················· | ···················· | ···················· | ···················· |
| G | 2465 hCoV-19/USA/OR-OHSU-1889/2020\|EPI_ISL_526060\|2020- | ···················· | ···················· | ···················· | ···················· | ···················· | ···················· |
| G | 2466 hCoV-19/USA/OR-OHSU-1893/2020\|EPI_ISL_526061\|2020- | ···················· | ···················· | ···················· | ···················· | ···················· | ···················· |
| G | 2467 hCoV-19/USA/OR-OHSU-1895/2020\|EPI_ISL_526062\|2020- | ···················· | ···················· | ···················· | ···················· | ···················· | ···················· |
| G | 2468 hCoV-19/USA/OR-OHSU-1897/2020\|EPI_ISL_526063\|2020- | ···················· | ···················· | ···················· | ···················· | ···················· | ···················· |
| G | 2469 hCoV-19/USA/OR-OHSU-1903/2020\|EPI_ISL_526064\|2020- | ···················· | ···················· | ···················· | ···················· | ···················· | ···················· |
| G | 2470 hCoV-19/USA/OR-OHSU-1905/2020\|EPI_ISL_526065\|2020- | ···················· | ·A·················· | ···················· | ···················· | ···················· | ···················· |
| G | 2471 hCoV-19/USA/OR-OHSU-1907/2020\|EPI_ISL_526066\|2020- | ···················· | ···················· | ···················· | ···················· | ···················· | ···················· |
| G | 2472 hCoV-19/USA/OR-OHSU-1911/2020\|EPI_ISL_526067\|2020- | ···················· | ···················· | ···················· | ···················· | ···················· | ···················· |
| G | 2473 hCoV-19/USA/OR-OHSU-1913/2020\|EPI_ISL_526068\|2020- | ···················· | ···················· | ···················· | ···················· | ···················· | ···················· |
| G | 2474 hCoV-19/USA/OR-OHSU-1915/2020\|EPI_ISL_526069\|2020- | ···················· | ·A·················· | ···················· | ···················· | ···················· | ···················· |
| G | 2475 hCoV-19/USA/OR-OHSU-1923/2020\|EPI_ISL_526071\|2020- | ···················· | ·A·················· | ···················· | ···················· | ···················· | ···················· |
| G | 2476 hCoV-19/USA/OR-OHSU-1925/2020\|EPI_ISL_526072\|2020- | ···················· | ·A·················· | ···················· | ···················· | ···················· | ···················· |
| G | 2477 hCoV-19/USA/OR-OHSU-1929/2020\|EPI_ISL_526073\|2020- | ···················· | ···················· | ···················· | ···················· | ···················· | ···················· |
| G | 2478 hCoV-19/USA/OR-OHSU-1935/2020\|EPI_ISL_526074\|2020- | ···················· | ·A·················· | ···················· | ···················· | ···················· | ···················· |
| G | 2479 hCoV-19/USA/OR-OHSU-1937/2020\|EPI_ISL_526075\|2020- | ···················· | ···················· | ···················· | ···················· | ···················· | ···················· |
| G | 2480 hCoV-19/USA/OR-OHSU-1945/2020\|EPI_ISL_526076\|2020- | ···················· | ·A·················· | ···················· | ···················· | ···················· | ···················· |
| G | 2481 hCoV-19/USA/OR-OHSU-1959/2020\|EPI_ISL_526078\|2020- | ···················· | ·A·················· | ···················· | ···················· | ···················· | ···················· |
| G | 2482 hCoV-19/USA/OR-OHSU-1971/2020\|EPI_ISL_526079\|2020- | ···················· | ···················· | ···················· | ···················· | ···················· | ···················· |
| G | 2483 hCoV-19/USA/OR-OHSU-1973/2020\|EPI_ISL_526080\|2020- | ···················· | ···················· | ···················· | ···················· | ···················· | ···················· |
| G | 2484 hCoV-19/USA/OR-OHSU-1981/2020\|EPI_ISL_526082\|2020- | ···················· | ···················· | ···················· | ···················· | ···················· | ···················· |
| G | 2485 hCoV-19/USA/OR-OHSU-1999/2020\|EPI_ISL_526085\|2020- | ···················· | ···················· | ···················· | ···················· | ···················· | ···················· |
| G | 2486 hCoV-19/South Korea/KCDC2766/2020\|EPI_ISL_526700\|2 | ···················· | ···················· | ···················· | ···················· | ···················· | ···················· |
| G | 2487 hCoV-19/South Korea/KCDC2767/2020\|EPI_ISL_526701\|2 | ···················· | ···················· | ···················· | ···················· | ···················· | ···················· |
| G | 2488 hCoV-19/South Korea/KCDC2768/2020\|EPI_ISL_526702\|2 | ···················· | ···················· | ···················· | ···················· | ···················· | ···················· |
| G | 2489 hCoV-19/South Korea/KCDC2769/2020\|EPI_ISL_526703\|2 | ···················· | ···················· | ···················· | ···················· | ···················· | ···················· |
| G | 2490 hCoV-19/England/201790460/2020\|EPI_ISL_526766\|2020 | ···················· | ···················· | ···················· | ···················· | ···················· | ···················· |
| G | 2491 hCoV-19/England/201790463/2020\|EPI_ISL_526767\|2020 | ···················· | ···················· | ···················· | ···················· | ···················· | ···················· |
| G | 2492 hCoV-19/England/201790473/2020\|EPI_ISL_526770\|2020 | ···················· | ···················· | ···················· | ···················· | ···················· | ···················· |
| G | 2493 hCoV-19/England/201861242/2020\|EPI_ISL_526772\|2020 | ···················· | ···················· | ···················· | ···················· | ···················· | ···················· |
| G | 2494 hCoV-19/England/201890916/2020\|EPI_ISL_526773\|2020 | ···················· | ···················· | ···················· | ···················· | ···················· | ···················· |
| G | 2495 hCoV-19/USA/VA-DCLS-0467/2020\|EPI_ISL_526775\|2020- | ···················· | ···················· | ···················· | ···················· | ···················· | ·········A·········· |
| G | 2496 hCoV-19/USA/VA-DCLS-0468/2020\|EPI_ISL_526776\|2020- | ···················· | ···················· | ···················· | ···················· | ···················· | ·········A·········· |
| G | 2497 hCoV-19/USA/VA-DCLS-0469/2020\|EPI_ISL_526777\|2020- | ···················· | ···················· | ···················· | ···················· | ···················· | ···················· |
| G | 2498 hCoV-19/USA/VA-DCLS-0470/2020\|EPI_ISL_526778\|2020- | ···················· | ···················· | ···················· | ···················· | ···················· | ···················· |
| G | 2499 hCoV-19/USA/VA-DCLS-0471/2020\|EPI_ISL_526779\|2020- | ···················· | ···················· | ···················· | ···················· | ···················· | ···················· |
| G | 2500 hCoV-19/USA/VA-DCLS-0472/2020\|EPI_ISL_526780\|2020- | ···················· | ···················· | ···················· | ···················· | ···················· | ···················· |
| G | 2501 hCoV-19/USA/VA-DCLS-0473/2020\|EPI_ISL_526781\|2020- | ···················· | ···················· | ···················· | ···················· | ···················· | ···················· |
| G | 2502 hCoV-19/USA/VA-DCLS-0474/2020\|EPI_ISL_526782\|2020- | ···················· | ···················· | ···················· | ···················· | ···················· | ···················· |
| G | 2503 hCoV-19/USA/VA-DCLS-0475/2020\|EPI_ISL_526783\|2020- | ···················· | ···················· | ···················· | ···················· | ···················· | ···················· |
| G | 2504 hCoV-19/USA/VA-DCLS-0476/2020\|EPI_ISL_526784\|2020- | ···················· | ···················· | ···················· | ···················· | ···················· | ···················· |

| | | | | | | | |
|---|---|---|---|---|---|---|---|
| G | 2505 hCoV-19/USA/VA-DCLS-0477/2020\|EPI_ISL_526785\|2020- | ···················· | ···················· | ····················· | ···················· | ···················· | ···················· |
| G | 2506 hCoV-19/USA/VA-DCLS-0478/2020\|EPI_ISL_526786\|2020- | ···················· | ···················· | ····················· | ···················· | ···················· | ···················· |
| G | 2507 hCoV-19/USA/VA-DCLS-0479/2020\|EPI_ISL_526787\|2020- | ···················· | ···················· | ····················· | ···················· | ···················· | ···················· |
| G | 2508 hCoV-19/USA/VA-DCLS-0480/2020\|EPI_ISL_526788\|2020- | ···················· | ···················· | ····················· | ···················· | ···················· | ···················· |
| G | 2509 hCoV-19/USA/VA-DCLS-0481/2020\|EPI_ISL_526789\|2020- | ···················· | ···················· | ····················· | ···················· | ···················· | ···················· |
| G | 2510 hCoV-19/Singapore/851/2020\|EPI_ISL_527360\|2020-08- | ···················· | ···················· | ····················· | ···················· | ···················· | ···················· |
| G | 2511 hCoV-19/Singapore/852/2020\|EPI_ISL_527361\|2020-08- | ···················· | ···················· | ····················· | ···················· | ···················· | ···················· |
| G | 2512 hCoV-19/Singapore/853/2020\|EPI_ISL_527362\|2020-08- | ···················· | ···················· | ····················· | ···················· | ···················· | ···················· |
| G | 2513 hCoV-19/Singapore/854/2020\|EPI_ISL_527363\|2020-08- | ···················· | ···················· | ····················· | ···················· | ···················· | ···················· |
| G | 2514 hCoV-19/Singapore/855/2020\|EPI_ISL_527364\|2020-08- | ···················· | ···················· | ····················· | ···················· | ···················· | ···················· |
| G | 2515 hCoV-19/Singapore/856/2020\|EPI_ISL_527365\|2020-08- | ···················· | ···················· | ····················· | ···················· | ···················· | ···················· |
| G | 2516 hCoV-19/Singapore/857/2020\|EPI_ISL_527366\|2020-08- | ···················· | ···················· | ····················· | ···················· | ···················· | ···················· |
| G | 2517 hCoV-19/Singapore/858/2020\|EPI_ISL_527367\|2020-08- | ···················· | ···················· | ····················· | ···················· | ···················· | ···················· |
| G | 2518 hCoV-19/Singapore/859/2020\|EPI_ISL_527368\|2020-08- | ···················· | ···················· | ····················· | ···················· | ···················· | ···················· |
| G | 2519 hCoV-19/Singapore/861/2020\|EPI_ISL_527370\|2020-04- | ···················· | ···················· | ····················· | ···················· | ···················· | ···················· |
| G | 2520 hCoV-19/Singapore/862/2020\|EPI_ISL_527371\|2020-04- | ···················· | ···················· | ····················· | ···················· | ···················· | ···················· |
| G | 2521 hCoV-19/Singapore/863/2020\|EPI_ISL_527372\|2020-04- | ···················· | ···················· | ····················· | ···················· | ···················· | ···················· |
| G | 2522 hCoV-19/Singapore/864/2020\|EPI_ISL_527373\|2020-08- | ···················· | ···················· | ····················· | ···················· | ···················· | ···················· |
| G | 2523 hCoV-19/Singapore/865/2020\|EPI_ISL_527374\|2020-08- | ···················· | ···················· | ····················· | ···················· | ···················· | ···················· |
| G | 2524 hCoV-19/Singapore/866/2020\|EPI_ISL_527375\|2020-04- | ···················· | ···················· | ····················· | ···················· | ···················· | ···················· |
| G | 2525 hCoV-19/Singapore/867/2020\|EPI_ISL_527376\|2020-04- | ···················· | ···················· | ····················· | ···················· | ···················· | ···················· |
| G | 2526 hCoV-19/Singapore/868/2020\|EPI_ISL_527377\|2020-04- | ···················· | ···················· | ····················· | ···················· | ···················· | ···················· |
| G | 2527 hCoV-19/Singapore/870/2020\|EPI_ISL_527379\|2020-08- | ···················· | ···················· | ····················· | ···················· | ···················· | ···················· |
| G | 2528 hCoV-19/Italy/APU-UniMI-4663/2020\|EPI_ISL_527380\|2 | ···················· | ···················· | ····················· | ···················· | ···················· | ···················· |
| G | 2529 hCoV-19/USA/PR-B2DR/2020\|EPI_ISL_527382\|2020-08-04 | ···················· | ···················· | ····················· | ···················· | ···················· | ···················· |
| G | 2530 hCoV-19/USA/PR-B2FH/2020\|EPI_ISL_527383\|2020-08-04 | ···················· | ···················· | ····················· | ···················· | ···················· | ···················· |
| G | 2531 hCoV-19/USA/PR-B2K5/2020\|EPI_ISL_527384\|2020-08-04 | ···················· | ···················· | ····················· | ···················· | ···················· | ···················· |
| G | 2532 hCoV-19/USA/PR-B35A/2020\|EPI_ISL_527385\|2020-08-05 | ···················· | ···················· | ····················· | ···················· | ···················· | ···················· |
| G | 2533 hCoV-19/USA/PR-B35C/2020\|EPI_ISL_527386\|2020-08-05 | ···················· | ···················· | ····················· | ···················· | ···················· | ···················· |
| G | 2534 hCoV-19/USA/PR-B40A/2020\|EPI_ISL_527387\|2020-08-06 | ···················· | ···················· | ····················· | ···················· | ···················· | ···················· |
| G | 2535 hCoV-19/USA/PR-B41C/2020\|EPI_ISL_527388\|2020-08-06 | ···················· | ···················· | ····················· | ···················· | ···················· | ···················· |
| G | 2536 hCoV-19/USA/PR-B5C5/2020\|EPI_ISL_527389\|2020-08-08 | ···················· | ···················· | ····················· | ···················· | ···················· | ···················· |
| G | 2537 hCoV-19/USA/PR-B5X4/2020\|EPI_ISL_527390\|2020-08-11 | ···················· | ···················· | ····················· | ···················· | ···················· | ···················· |
| G | 2538 hCoV-19/USA/PR-B5XH/2020\|EPI_ISL_527391\|2020-08-11 | ···················· | ···················· | ····················· | ···················· | ···················· | ···················· |
| G | 2539 hCoV-19/USA/PR-B5XJ/2020\|EPI_ISL_527392\|2020-08-11 | ···················· | ···················· | ····················· | ···················· | ···················· | ···················· |
| G | 2540 hCoV-19/USA/PR-B5XS/2020\|EPI_ISL_527393\|2020-08-11 | ···················· | ···················· | ····················· | ···················· | ···················· | ···················· |
| G | 2541 hCoV-19/USA/PR-B6BV/2020\|EPI_ISL_527394\|2020-08-11 | ···················· | ···················· | ····················· | ···················· | ···················· | ···················· |
| G | 2542 hCoV-19/USA/PR-B6CA/2020\|EPI_ISL_527395\|2020-08-11 | ···················· | ···················· | ····················· | ···················· | ···················· | ···················· |
| G | 2543 hCoV-19/USA/PR-B6CS/2020\|EPI_ISL_527396\|2020-08-11 | ···················· | ·A·················· | ····················· | ···················· | ···················· | ···················· |
| G | 2544 hCoV-19/USA/PR-B6ZT/2020\|EPI_ISL_527397\|2020-08-12 | ···················· | ···················· | ····················· | ···················· | ···················· | ···················· |
| G | 2545 hCoV-19/USA/PR-B74P/2020\|EPI_ISL_527398\|2020-08-12 | ···················· | ···················· | ····················· | ···················· | ···················· | ···················· |
| G | 2546 hCoV-19/USA/PR-B75X/2020\|EPI_ISL_527399\|2020-08-12 | ···················· | ···················· | ····················· | ···················· | ···················· | ···················· |
| G | 2547 hCoV-19/United Arab Emirates/16B/2020\|EPI_ISL_5286 | ···················· | ···················· | ····················· | ···················· | ···················· | ···················· |
| G | 2548 hCoV-19/United Arab Emirates/195/2020\|EPI_ISL_5286 | ···················· | ···················· | ····················· | ···················· | ···················· | ···················· |
| G | 2549 hCoV-19/United Arab Emirates/2/2020\|EPI_ISL_528692 | ···················· | ···················· | ····················· | ···················· | ···················· | ···················· |
| G | 2550 hCoV-19/United Arab Emirates/21R/2020\|EPI_ISL_5286 | ···················· | ···················· | ····················· | ···················· | ···················· | ···················· |
| G | 2551 hCoV-19/United Arab Emirates/306/2020\|EPI_ISL_5286 | ···················· | ···················· | ····················· | ···················· | ···················· | ···················· |
| G | 2552 hCoV-19/United Arab Emirates/310/2020\|EPI_ISL_5286 | ···················· | ···················· | ····················· | ···················· | ···················· | ···················· |
| G | 2553 hCoV-19/United Arab Emirates/313/2020\|EPI_ISL_5286 | ···················· | ···················· | ····················· | ···················· | ···················· | ···················· |
| G | 2554 hCoV-19/United Arab Emirates/31B/2020\|EPI_ISL_5286 | ···················· | ···················· | ····················· | ···················· | ···················· | ···················· |
| G | 2555 hCoV-19/United Arab Emirates/38/2020\|EPI_ISL_52869 | ···················· | ···················· | ····················· | ···················· | ···················· | ···················· |
| G | 2556 hCoV-19/United Arab Emirates/42/2020\|EPI_ISL_52869 | ···················· | ···················· | ····················· | ···················· | ···················· | ···················· |
| G | 2557 hCoV-19/Japan/TK-YE63751/2020\|EPI_ISL_582144\|2020- | ···················· | ···················· | ····················· | ···················· | ···················· | ···················· |
| G | 2558 hCoV-19/Japan/TK-YE63751d/2020\|EPI_ISL_582145\|2020 | ···················· | ···················· | ····················· | ···················· | ···················· | ···················· |
| G | 2559 hCoV-19/Japan/TK-YE64688/2020\|EPI_ISL_582146\|2020- | ···················· | ···················· | ····················· | ···················· | ···················· | ···················· |
| G | 2560 hCoV-19/Japan/TK-YE64688d/2020\|EPI_ISL_582147\|2020 | ···················· | ···················· | ····················· | ···················· | ···················· | ···················· |
| G | 2561 hCoV-19/England/LIVE-A750C/2020\|EPI_ISL_517031\|202 | ···················· | ···················· | ····················· | ···················· | ···················· | ···················· |
| G | 2565 hCoV-19/England/LIVE-A7566/2020\|EPI_ISL_517037\|202 | ···················· | ···················· | ····················· | ···················· | ···················· | ···················· |
| G | 2566 hCoV-19/Australia/VIC5526/2020\|EPI_ISL_518360\|2020 | ···················· | ···················· | ····················· | ···················· | ···················· | ···················· |
| G | 2567 hCoV-19/Australia/VIC5528/2020\|EPI_ISL_518361\|2020 | ···················· | ···················· | ····················· | ···················· | ···················· | ···················· |
| G | 2568 hCoV-19/Australia/VIC5529/2020\|EPI_ISL_518362\|2020 | ···················· | ···················· | ····················· | ···················· | ···················· | ···················· |
| G | 2569 hCoV-19/Australia/VIC5531/2020\|EPI_ISL_518363\|2020 | ···················· | ···················· | ····················· | ···················· | ···················· | ···················· |
| G | 2570 hCoV-19/Australia/VIC5532/2020\|EPI_ISL_518364\|2020 | ···················· | ···················· | ····················· | ···················· | ···················· | ···················· |
| G | 2571 hCoV-19/Australia/VIC5534/2020\|EPI_ISL_518365\|2020 | ···················· | ···················· | ····················· | ···················· | ···················· | ···················· |
| G | 2572 hCoV-19/Australia/VIC5536/2020\|EPI_ISL_518367\|2020 | ···················· | ···················· | ····················· | ···················· | ···················· | ···················· |
| G | 2573 hCoV-19/Australia/VIC5537/2020\|EPI_ISL_518368\|2020 | ···················· | ···················· | ····················· | ···················· | ···················· | ···················· |
| G | 2574 hCoV-19/Australia/VIC4272/2020\|EPI_ISL_519600\|2020 | ···················· | ···················· | ····················· | ···················· | ···················· | ···················· |
| G | 2575 hCoV-19/Australia/VIC4273/2020\|EPI_ISL_519601\|2020 | ···················· | ···················· | ····················· | ···················· | ···················· | ···················· |
| G | 2576 hCoV-19/Australia/VIC4274/2020\|EPI_ISL_519602\|2020 | ···················· | ···················· | ····················· | ···················· | ···················· | ···················· |
| G | 2577 hCoV-19/Australia/VIC4275/2020\|EPI_ISL_519603\|2020 | ···················· | ···················· | ····················· | ···················· | ···················· | ···················· |
| G | 2578 hCoV-19/Australia/VIC4276/2020\|EPI_ISL_519604\|2020 | ···················· | ···················· | ····················· | ···················· | ···················· | ···················· |
| G | 2579 hCoV-19/Australia/VIC4277/2020\|EPI_ISL_519605\|2020 | ···················· | ···················· | ····················· | ···················· | ···················· | ···················· |
| G | 2580 hCoV-19/Australia/VIC4278/2020\|EPI_ISL_519606\|2020 | ···················· | ···················· | ····················· | ···················· | ···················· | ···················· |
| G | 2581 hCoV-19/Australia/VIC4279/2020\|EPI_ISL_519607\|2020 | ···················· | ···················· | ····················· | ···················· | ···················· | ···················· |
| G | 2582 hCoV-19/Australia/VIC4280/2020\|EPI_ISL_519608\|2020 | ···················· | ···················· | ····················· | ···················· | ···················· | ···················· |
| G | 2583 hCoV-19/Australia/VIC4281/2020\|EPI_ISL_519609\|2020 | ···················· | ···················· | ····················· | ···················· | ···················· | ···················· |
| G | 2584 hCoV-19/Australia/VIC4282/2020\|EPI_ISL_519610\|2020 | ···················· | ···················· | ····················· | ···················· | ···················· | ···················· |
| G | 2585 hCoV-19/Australia/VIC4283/2020\|EPI_ISL_519611\|2020 | ···················· | ···················· | ····················· | ···················· | ···················· | ···················· |
| G | 2586 hCoV-19/Australia/VIC4284/2020\|EPI_ISL_519612\|2020 | ···················· | ···················· | ····················· | ···················· | ···················· | ···················· |
| G | 2587 hCoV-19/Australia/VIC4285/2020\|EPI_ISL_519613\|2020 | ···················· | ···················· | ····················· | ···················· | ···················· | ···················· |

| | | | | | | | |
|---|---|---|---|---|---|---|---|
| G | 2588 hCoV-19/Australia/VIC4286/2020\|EPI_ISL_519614\|2020 | ···················· | ···················· | ···················· | ···················· | ···················· | ···················· |
| G | 2589 hCoV-19/Australia/VIC4287/2020\|EPI_ISL_519615\|2020 | ···················· | ···················· | ···················· | ···················· | ···················· | ···················· |
| G | 2590 hCoV-19/Australia/VIC4288/2020\|EPI_ISL_519616\|2020 | ···················· | ···················· | ···················· | ···················· | ···················· | ···················· |
| G | 2591 hCoV-19/Australia/VIC4320/2020\|EPI_ISL_519617\|2020 | ···················· | ···················· | ···················· | ···················· | ···················· | ···················· |
| G | 2592 hCoV-19/Australia/VIC4321/2020\|EPI_ISL_519618\|2020 | ···················· | ···················· | ···················· | ···················· | ···················· | ···················· |
| G | 2593 hCoV-19/Australia/VIC4322/2020\|EPI_ISL_519619\|2020 | ···················· | ···················· | ···················· | ···················· | ···················· | ···················· |
| G | 2594 hCoV-19/Australia/VIC4395/2020\|EPI_ISL_519691\|2020 | ···················· | ···················· | ···················· | ···················· | ···················· | ···················· |
| G | 2595 hCoV-19/Australia/VIC4396/2020\|EPI_ISL_519692\|2020 | ···················· | ···················· | ···················· | ···················· | ···················· | ···················· |
| G | 2596 hCoV-19/Australia/VIC4397/2020\|EPI_ISL_519693\|2020 | ···················· | ···················· | ···················· | ···················· | ···················· | ···················· |
| G | 2597 hCoV-19/Australia/VIC4398/2020\|EPI_ISL_519694\|2020 | ···················· | ···················· | ···················· | ···················· | ···················· | ···················· |
| G | 2598 hCoV-19/Australia/VIC4399/2020\|EPI_ISL_519695\|2020 | ···················· | ···················· | ···················· | ···················· | ···················· | ···················· |
| G | 2599 hCoV-19/Australia/VIC4400/2020\|EPI_ISL_519696\|2020 | ···················· | ···················· | ···················· | ···················· | ···················· | ···················· |
| G | 2600 hCoV-19/Australia/VIC4401/2020\|EPI_ISL_519697\|2020 | ···················· | ···················· | ···················· | ···················· | ···················· | ···················· |
| G | 2601 hCoV-19/Australia/VIC4402/2020\|EPI_ISL_519698\|2020 | ···················· | ···················· | ···················· | ···················· | ···················· | ···················· |
| G | 2602 hCoV-19/United Arab Emirates/L2005036714/2020\|EPI_ | ···················· | ···················· | ···················· | ···················· | ···················· | ···················· |
| G | 2603 hCoV-19/United Arab Emirates/L2006030187/2020\|EPI_ | ···················· | ···················· | ···················· | ···················· | ···················· | ···················· |
| G | 2604 hCoV-19/United Arab Emirates/L2006030255/2020\|EPI_ | ···················· | ···················· | ···················· | ···················· | ···················· | ···················· |
| G | 2605 hCoV-19/United Arab Emirates/L2006032095/2020\|EPI_ | ···················· | ···················· | ···················· | ···················· | ···················· | ···················· |
| G | 2606 hCoV-19/United Arab Emirates/L3280050877/2020\|EPI_ | ···················· | ···················· | ···················· | ···················· | ···················· | ···················· |
| G | 2607 hCoV-19/United Arab Emirates/L3290161092/2020\|EPI_ | ···················· | ···················· | ···················· | ···················· | ···················· | ···················· |
| G | 2608 hCoV-19/United Arab Emirates/L3290162047/2020\|EPI_ | ···················· | ···················· | ···················· | ···················· | ···················· | ···················· |
| G | 2609 hCoV-19/United Arab Emirates/L3290165435/2020\|EPI_ | ···················· | ···················· | ···················· | ···················· | ···················· | ···················· |
| G | 2610 hCoV-19/United Arab Emirates/L3290165790/2020\|EPI_ | ···················· | ···················· | ···················· | ···················· | ···················· | ···················· |
| G | 2611 hCoV-19/United Arab Emirates/L3290166145/2020\|EPI_ | ···················· | ···················· | ···················· | ···················· | ···················· | ···················· |
| G | 2612 hCoV-19/United Arab Emirates/L3290201241/2020\|EPI_ | ···················· | ···················· | ···················· | ···················· | ···················· | ···················· |
| G | 2613 hCoV-19/Australia/VIC1994/2020\|EPI_ISL_521900\|2020 | ···················· | ···················· | ···················· | ···················· | ···················· | ···················· |
| G | 2614 hCoV-19/Australia/VIC1995/2020\|EPI_ISL_521901\|2020 | ···················· | ···················· | ···················· | ···················· | ···················· | ···················· |
| G | 2615 hCoV-19/Australia/VIC1996/2020\|EPI_ISL_521902\|2020 | ···················· | ···················· | ···················· | ···················· | ···················· | ···················· |
| G | 2616 hCoV-19/Australia/VIC2006/2020\|EPI_ISL_521903\|2020 | ···················· | ···················· | ···················· | ···················· | ···················· | ···················· |
| G | 2618 hCoV-19/Australia/VIC2045/2020\|EPI_ISL_521906\|2020 | ···················· | ···················· | ···················· | ···················· | ···················· | ···················· |
| G | 2619 hCoV-19/Australia/VIC2046/2020\|EPI_ISL_521907\|2020 | ···················· | ···················· | ···················· | ···················· | ···················· | ···················· |
| G | 2620 hCoV-19/Australia/VIC2049/2020\|EPI_ISL_521909\|2020 | ···················· | ···················· | ···················· | ···················· | ···················· | ···················· |
| G | 2621 hCoV-19/Australia/VIC2075/2020\|EPI_ISL_521910\|2020 | ···················· | ···················· | ···················· | ···················· | ···················· | ···················· |
| G | 2622 hCoV-19/Australia/VIC2081/2020\|EPI_ISL_521912\|2020 | ···················· | ···················· | ···················· | ···················· | ···················· | ···················· |
| G | 2623 hCoV-19/Australia/VIC2082/2020\|EPI_ISL_521913\|2020 | ···················· | ···················· | ···················· | ···················· | ···················· | ···················· |
| G | 2624 hCoV-19/Australia/VIC2083/2020\|EPI_ISL_521914\|2020 | ···················· | ···················· | ···················· | ···················· | ···················· | ···················· |
| G | 2625 hCoV-19/Australia/VIC2147/2020\|EPI_ISL_521916\|2020 | ···················· | ···················· | ···················· | ···················· | ···················· | ···················· |
| G | 2626 hCoV-19/Australia/VIC2148/2020\|EPI_ISL_521917\|2020 | ···················· | ···················· | ···················· | ···················· | ···················· | ···················· |
| G | 2627 hCoV-19/Australia/VIC2169/2020\|EPI_ISL_521918\|2020 | ···················· | ···················· | ···················· | ···················· | ···················· | ···················· |
| G | 2628 hCoV-19/Australia/VIC2170/2020\|EPI_ISL_521919\|2020 | ···················· | ···················· | ···················· | ···················· | ·NN················· | ···················· |
| G | 2630 hCoV-19/Australia/VIC2173/2020\|EPI_ISL_521922\|2020 | ···················· | ···················· | ···················· | ···················· | ···················· | ···················· |
| G | 2631 hCoV-19/Australia/VIC2175/2020\|EPI_ISL_521924\|2020 | ···················· | ···················· | ···················· | ···················· | ···················· | ···················· |
| G | 2632 hCoV-19/Australia/VIC2176/2020\|EPI_ISL_521925\|2020 | ···················· | ···················· | ···················· | ···················· | ···················· | ···················· |
| G | 2633 hCoV-19/Australia/VIC2181/2020\|EPI_ISL_521927\|2020 | ···················· | ···················· | ···················· | ···················· | ···················· | ···················· |
| G | 2634 hCoV-19/Australia/VIC2182/2020\|EPI_ISL_521928\|2020 | ···················· | ···················· | ···················· | ···················· | ···················· | ···················· |
| G | 2635 hCoV-19/Australia/VIC2183/2020\|EPI_ISL_521929\|2020 | ···················· | ···················· | ···················· | ···················· | ···················· | ···················· |
| G | 2636 hCoV-19/USA/UT-08304/2020\|EPI_ISL_524100\|2020-05-1 | ···················· | ···················· | ···················· | ···················· | ···················· | ···················· |
| G | 2637 hCoV-19/USA/UT-08305/2020\|EPI_ISL_524101\|2020-05-2 | ···················· | ···················· | ···················· | ···················· | ···················· | ···················· |
| G | 2638 hCoV-19/USA/UT-08306/2020\|EPI_ISL_524102\|2020-05-2 | ···················· | ···················· | ···················· | ···················· | ···················· | ···················· |
| G | 2639 hCoV-19/USA/UT-08309/2020\|EPI_ISL_524105\|2020-05-2 | ···················· | ···················· | ···················· | ···················· | ···················· | ···················· |
| G | 2640 hCoV-19/USA/UT-08311/2020\|EPI_ISL_524107\|2020-05-2 | ···················· | ···················· | ···················· | ···················· | ···················· | ···················· |
| G | 2641 hCoV-19/USA/UT-08342/2020\|EPI_ISL_524130\|2020-05-1 | ···················· | ···················· | ···················· | ···················· | ···················· | ···················· |
| G | 2642 hCoV-19/USA/UT-08346/2020\|EPI_ISL_524133\|2020-05-1 | ···················· | ···················· | ···················· | ···················· | ···················· | ···················· |
| G | 2643 hCoV-19/USA/UT-08348/2020\|EPI_ISL_524134\|2020-05-1 | ···················· | ·A·················· | ···················· | ···················· | ···················· | ···················· |
| G | 2644 hCoV-19/USA/UT-08349/2020\|EPI_ISL_524135\|2020-05-1 | ···················· | ···················· | ···················· | ···················· | ···················· | ···················· |
| G | 2645 hCoV-19/USA/UT-08350/2020\|EPI_ISL_524136\|2020-05-1 | ···················· | ···················· | ···················· | ···················· | ···················· | ···················· |
| G | 2646 hCoV-19/USA/UT-08353/2020\|EPI_ISL_524138\|2020-05-1 | ···················· | ···················· | ···················· | ···················· | ···················· | ···················· |
| G | 2647 hCoV-19/USA/UT-08354/2020\|EPI_ISL_524139\|2020-05-2 | ···················· | ···················· | ···················· | ···················· | ···················· | ···················· |
| G | 2648 hCoV-19/USA/UT-08358/2020\|EPI_ISL_524141\|2020-05-1 | ···················· | ···················· | ···················· | ···················· | ···················· | ···················· |
| G | 2649 hCoV-19/USA/UT-08359/2020\|EPI_ISL_524142\|2020-05-1 | ···················· | ···················· | ···················· | ···················· | ···················· | ···················· |
| G | 2650 hCoV-19/USA/UT-08360/2020\|EPI_ISL_524143\|2020-05-1 | ···················· | ···················· | ···················· | ···················· | ···················· | ···················· |
| G | 2651 hCoV-19/USA/UT-08362/2020\|EPI_ISL_524145\|2020-05-2 | ···················· | ···················· | ···················· | ···················· | ···················· | ···················· |
| G | 2652 hCoV-19/USA/UT-08364/2020\|EPI_ISL_524147\|2020-05-2 | ···················· | ···················· | ···················· | ···················· | ···················· | ···················· |
| G | 2653 hCoV-19/USA/UT-08365/2020\|EPI_ISL_524148\|2020-05-1 | ···················· | ···················· | ···················· | ···················· | ···················· | ···················· |
| G | 2654 hCoV-19/USA/UT-08366/2020\|EPI_ISL_524149\|2020-05-2 | ···················· | ···················· | ···················· | ···················· | ···················· | ···················· |
| G | 2655 hCoV-19/USA/UT-08368/2020\|EPI_ISL_524151\|2020-05-1 | ···················· | ···················· | ···················· | ···················· | ···················· | ···················· |
| G | 2656 hCoV-19/USA/UT-08369/2020\|EPI_ISL_524152\|2020-05-1 | ···················· | ···················· | ···················· | ···················· | ···················· | ···················· |
| G | 2657 hCoV-19/USA/UT-08371/2020\|EPI_ISL_524154\|2020-05-1 | ···················· | ···················· | ···················· | ···················· | ···················· | ···················· |
| G | 2658 hCoV-19/USA/UT-08372/2020\|EPI_ISL_524155\|2020-05-1 | ···················· | ···················· | ···················· | ···················· | ···················· | ···················· |
| G | 2659 hCoV-19/USA/UT-08374/2020\|EPI_ISL_524157\|2020-05-2 | ···················· | ···················· | ···················· | ···················· | ···················· | ···················· |
| G | 2660 hCoV-19/USA/UT-08378/2020\|EPI_ISL_524161\|2020-05-2 | ···················· | ···················· | ···················· | ···················· | ···················· | ···················· |
| G | 2661 hCoV-19/USA/UT-08379/2020\|EPI_ISL_524162\|2020-05-1 | ···················· | ···················· | ···················· | ···················· | ···················· | ···················· |
| G | 2662 hCoV-19/USA/UT-08380/2020\|EPI_ISL_524163\|2020-05-1 | ···················· | ···················· | ···················· | ···················· | ···················· | ···················· |
| G | 2663 hCoV-19/USA/UT-08381/2020\|EPI_ISL_524164\|2020-05-1 | ···················· | ···················· | ···················· | ···················· | ···················· | ···················· |
| G | 2664 hCoV-19/USA/UT-08382/2020\|EPI_ISL_524165\|2020-05-1 | ···················· | ···················· | ···················· | ···················· | ···················· | ···················· |
| G | 2665 hCoV-19/USA/UT-08383/2020\|EPI_ISL_524166\|2020-05-1 | ···················· | ···················· | ···················· | ···················· | ···················· | ···················· |
| G | 2666 hCoV-19/USA/UT-08384/2020\|EPI_ISL_524167\|2020-05-2 | ···················· | ···················· | ···················· | ···················· | ···················· | ···················· |
| G | 2667 hCoV-19/USA/UT-08387/2020\|EPI_ISL_524170\|2020-05-2 | ···················· | ···················· | ···················· | ···················· | ···················· | ···················· |
| G | 2668 hCoV-19/USA/UT-08389/2020\|EPI_ISL_524171\|2020-05-2 | ···················· | ···················· | ···················· | ···················· | ···················· | ···················· |
| G | 2669 hCoV-19/USA/UT-08391/2020\|EPI_ISL_524172\|2020-05-1 | ···················· | ···················· | ···················· | ···················· | ···················· | ···················· |

```
G  2670 hCoV-19/USA/UT-08392/2020|EPI_ISL_524173|2020-05-1  ···················· ···················· ···················· ···················· ···················· ····················
G  2671 hCoV-19/USA/UT-08393/2020|EPI_ISL_524174|2020-05-1  ···················· ···················· ···················· ···················· ···················· ····················
G  2672 hCoV-19/USA/UT-08395/2020|EPI_ISL_524175|2020-05-2  ···················· ···················· ···················· ···················· ···················· ····················
G  2673 hCoV-19/USA/UT-08396/2020|EPI_ISL_524176|2020-05-2  ···················· ···················· ···················· ···················· ···················· ····················
G  2674 hCoV-19/USA/UT-08398/2020|EPI_ISL_524178|2020-05-2  ···················· ···················· ···················· ···················· ···················· ····················
G  2675 hCoV-19/USA/UT-08399/2020|EPI_ISL_524179|2020-05-2  ···················· ···················· ···················· ···················· ···················· ····················
G  2676 hCoV-19/USA/UT-08400/2020|EPI_ISL_524180|2020-05-2  ···················· ···················· ···················· ···················· ···················· ····················
G  2677 hCoV-19/USA/UT-08402/2020|EPI_ISL_524181|2020-05-1  ···················· ···················· ···················· ···················· ···················· ····················
G  2678 hCoV-19/USA/UT-08403/2020|EPI_ISL_524182|2020-05-1  ···················· ···················· ···················· ···················· ···················· ····················
G  2679 hCoV-19/USA/UT-08404/2020|EPI_ISL_524183|2020-05-1  ···················· ···················· ···················· ···················· ···················· ····················
G  2680 hCoV-19/USA/UT-08406/2020|EPI_ISL_524184|2020-05-1  ···················· ···················· ···················· ···················· ···················· ····················
G  2681 hCoV-19/USA/UT-08408/2020|EPI_ISL_524186|2020-05-2  ···················· ···················· ···················· ···················· ···················· ····················
G  2682 hCoV-19/USA/UT-08409/2020|EPI_ISL_524187|2020-05-2  ···················· ···················· ···················· ···················· ···················· ····················
G  2683 hCoV-19/USA/UT-08410/2020|EPI_ISL_524188|2020-05-2  ···················· ···················· ···················· ···················· ···················· ····················
G  2684 hCoV-19/USA/UT-08412/2020|EPI_ISL_524190|2020-05-1  ···················· ···················· ···················· ···················· ···················· ····················
G  2685 hCoV-19/USA/UT-08413/2020|EPI_ISL_524191|2020-05-1  ···················· ···················· ···················· ···················· ···················· ····················
G  2686 hCoV-19/USA/UT-08414/2020|EPI_ISL_524192|2020-05-1  ···················· ···················· ···················· ···················· ···················· ····················
G  2687 hCoV-19/USA/UT-08415/2020|EPI_ISL_524193|2020-05-1  ···················· ···················· ···················· ···················· ···················· ····················
G  2688 hCoV-19/USA/UT-08416/2020|EPI_ISL_524194|2020-05-1  ···················· ···················· ···················· ···················· ···················· ····················
G  2689 hCoV-19/USA/UT-08418/2020|EPI_ISL_524196|2020-05-2  ···················· ···················· ···················· ···················· ···················· ····················
G  2690 hCoV-19/Ireland/KE-NVRL-72IRL00276/2020|EPI_ISL_52  ···················· ···················· ···················· ···················· ···················· ····················
G  2691 hCoV-19/Ireland/KE-NVRL-72IRL00278/2020|EPI_ISL_52  ···················· ···················· ···················· ···················· ···················· ····················
G  2692 hCoV-19/Ireland/OY-NVRL-72IRL00287/2020|EPI_ISL_52  ···················· ···················· ···················· ···················· ···················· ····················
G  2693 hCoV-19/Ireland/OY-NVRL-72IRL00288/2020|EPI_ISL_52  ···················· ···················· ···················· ···················· ···················· ····················
G  2694 hCoV-19/Ireland/KE-NVRL-72IRL00289/2020|EPI_ISL_52  ···················· ···················· ···················· ···················· ···················· ····················
G  2695 hCoV-19/Ireland/OY-NVRL-72IRL00290/2020|EPI_ISL_52  ···················· ···················· ···················· ···················· ···················· ····················
G  2696 hCoV-19/Ireland/OY-NVRL-72IRL00293/2020|EPI_ISL_52  ···················· ···················· ···················· ···················· ···················· ····················
G  2697 hCoV-19/Ireland/KE-NVRL-72IRL00302/2020|EPI_ISL_52  ···················· ···················· ···················· ···················· ···················· ····················
G  2698 hCoV-19/Ireland/OY-NVRL-72IRL00303/2020|EPI_ISL_52  ···················· ···················· ···················· ···················· ···················· ····················
G  2699 hCoV-19/Ireland/OY-NVRL-72IRL00304/2020|EPI_ISL_52  ···················· ···················· ···················· ···················· ···················· ····················
G  2700 hCoV-19/Ireland/OY-NVRL-72IRL00307/2020|EPI_ISL_52  ···················· ···················· ···················· ···················· ···················· ····················
G  2701 hCoV-19/Ireland/OY-NVRL-72IRL00318/2020|EPI_ISL_52  ···················· ···················· ···················· ···················· ···················· ····················
G  2702 hCoV-19/Ireland/OY-NVRL-72IRL00319/2020|EPI_ISL_52  ···················· ···················· ···················· ···················· ···················· ····················
G  2703 hCoV-19/Ireland/OY-NVRL-72IRL00323/2020|EPI_ISL_52  ···················· ···················· ···················· ···················· ···················· ····················
G  2704 hCoV-19/Ireland/KE-NVRL-72IRL00327/2020|EPI_ISL_52  ···················· ···················· ···················· ···················· ···················· ····················
G  2705 hCoV-19/Ireland/OY-NVRL-72IRL00330/2020|EPI_ISL_52  ···················· ···················· ···················· ···················· ···················· ····················
G  2706 hCoV-19/Ireland/OY-NVRL-72IRL00335/2020|EPI_ISL_52  ···················· ···················· ···················· ···················· ···················· ····················
G  2707 hCoV-19/Ireland/OY-NVRL-72IRL04484/2020|EPI_ISL_52  ···················· ···················· ···················· ···················· ···················· ····················
G  2708 hCoV-19/Ireland/KE-NVRL-72IRL06223/2020|EPI_ISL_52  ···················· ···················· ···················· ···················· ···················· ····················
G  2709 hCoV-19/India/GJ-GBRC-382a/2020|EPI_ISL_525419|202  ···················· ·A·················· ···················· ···················· ···················· ··A·················
G  2710 hCoV-19/India/GJ-GBRC-382b/2020|EPI_ISL_525420|202  ···················· ·A·················· ···················· ···················· ···················· ··A·················
G  2711 hCoV-19/India/GJ-GBRC-397a/2020|EPI_ISL_525421|202  ···················· ·A·················· ···················· ···················· ···················· ··A·················
G  2712 hCoV-19/India/GJ-GBRC-397b/2020|EPI_ISL_525422|202  ···················· ·A·················· ···················· ···················· ···················· ··A·················
G  2713 hCoV-19/Oman/16904/2020|EPI_ISL_525423|2020-04-12  ···················· ···················· ···················· ···················· ···················· ··A·················
G  2714 hCoV-19/Oman/B-2106/2020|EPI_ISL_525424|2020-04-21  ···················· ···················· ···················· ···················· ···················· ····················
G  2715 hCoV-19/Oman/RESP-20-4943/2020|EPI_ISL_525425|2020  ···················· ···················· ···················· ···················· ···················· ····················
G  2716 hCoV-19/Oman/RESP-20-B-15349/2020|EPI_ISL_525426|2  ···················· ···················· ···················· ···················· ···················· ····················
G  2717 hCoV-19/Oman/RESP-20-B-15491/2020|EPI_ISL_525427|2  ···················· ···················· ···················· ···················· ···················· ····················
G  2718 hCoV-19/Oman/RESP-20-B-15703/2020|EPI_ISL_525428|2  ···················· ···················· ···················· ···················· ···················· ····················
G  2719 hCoV-19/Oman/RESP-20-B-6588/2020|EPI_ISL_525429|20  ···················· ···················· ···················· ···················· ···················· ····················
G  2720 hCoV-19/Ecuador/USFQ-177/2020|EPI_ISL_525431|2020-  ···················· ···················· ···················· ···················· ···················· ····················
G  2721 hCoV-19/Ecuador/USFQ-178/2020|EPI_ISL_525432|2020-  ···················· ···················· ···················· ···················· ···················· ····················
G  2722 hCoV-19/Ecuador/USFQ-182/2020|EPI_ISL_525433|2020-  ···················· ···················· ···················· ···················· ···················· ····················
G  2723 hCoV-19/Ecuador/USFQ-183/2020|EPI_ISL_525434|2020-  ···················· ···················· ···················· ···················· ···················· ····················
G  2724 hCoV-19/Ecuador/USFQ-188/2020|EPI_ISL_525437|2020-  ···················· ···················· ···················· ···················· ···················· ····················
G  2725 hCoV-19/Ecuador/USFQ-MC01/2020|EPI_ISL_525438|2020  ···················· ···················· ···················· ···················· ···················· ····················
G  2726 hCoV-19/Oman/MA-RESP-20-B-16263/2020|EPI_ISL_52546  ···················· ···················· ···················· ···················· ···················· ····················
G  2727 hCoV-19/Dominican Republic/ICGEB_UNIBE022/2020|EPI  ···················· ···················· ···················· ···················· ···················· ····················
G  2728 hCoV-19/Dominican Republic/ICGEB_UNIBE032.2/2020|E  ···················· ·A·················· ···················· ···················· ···················· ····················
G  2729 hCoV-19/Dominican Republic/ICGEB_UNIBE032.7/2020|E  ···················· ···················· ···················· ···················· ···················· ····················
G  2730 hCoV-19/Sri Lanka/CDR142/2020|EPI_ISL_525474|2020-  ···················· ···················· ···················· ···················· ···················· ····················
G  2731 hCoV-19/Sri Lanka/CDR1885/2020|EPI_ISL_525476|2020  ···················· ···················· ···················· ···················· ···················· ····················
G  2732 hCoV-19/Sri Lanka/CDR194/2020|EPI_ISL_525478|2020-  ···················· ···················· ···················· ···················· ···················· ····················
G  2733 hCoV-19/Sri Lanka/CDR503/2020|EPI_ISL_525479|2020-  ···················· ···················· ···················· ···················· ···················· ····················
G  2734 hCoV-19/Sri Lanka/CDR-SL7066/2020|EPI_ISL_525481|2  ···················· ···················· ···················· T··················· ···················· ····················
G  2735 hCoV-19/Sri Lanka/CDR-KK57/2020|EPI_ISL_525486|202  ···················· ···················· ···················· ···················· ···················· ····················
G  2736 hCoV-19/Sri Lanka/CDR-KK230/2020|EPI_ISL_525489|20  ···················· ···················· ···················· ···················· ···················· ····················
G  2737 hCoV-19/Indonesia/JT-UGM-202538/2020|EPI_ISL_52549  ···················· ···················· ···················· ···················· ···················· ····················
G  2738 hCoV-19/Italy/FVG-ICGEB-S204/2020|EPI_ISL_525495|2  ···················· ···················· ···················· ···················· ···················· ····················
G  2739 hCoV-19/Italy/FVG-ICGEB-S205/2020|EPI_ISL_525496|2  ···················· ···················· ···················· ···················· ···················· ····················
G  2740 hCoV-19/South Korea/KCDC2770/2020|EPI_ISL_526704|2  ···················· ···················· ···················· ···················· ···················· ····················
G  2741 hCoV-19/South Korea/KCDC2771/2020|EPI_ISL_526705|2  ···················· ···················· ···················· ···················· ···················· ····················
G  2742 hCoV-19/South Korea/KCDC2772/2020|EPI_ISL_526706|2  ···················· ···················· ···················· ···················· ···················· ····················
G  2743 hCoV-19/South Korea/KCDC2773/2020|EPI_ISL_526707|2  ···················· ···················· ···················· ···················· ···················· ····················
G  2744 hCoV-19/South Korea/KCDC2774/2020|EPI_ISL_526708|2  ···················· ···················· ···················· ···················· ···················· ····················
G  2745 hCoV-19/South Korea/KCDC2775/2020|EPI_ISL_526709|2  ···················· ···················· ···················· ···················· ···················· ····················
G  2746 hCoV-19/South Korea/KCDC2776/2020|EPI_ISL_526710|2  ···················· ···················· ···················· ···················· ···················· ····················
G  2747 hCoV-19/South Korea/KCDC2777/2020|EPI_ISL_526711|2  ···················· ···················· ···················· ···················· ···················· ····················
G  2748 hCoV-19/South Korea/KCDC2778/2020|EPI_ISL_526712|2  ···················· ···················· ···················· ···················· ···················· ····················
G  2749 hCoV-19/South Korea/KCDC2779/2020|EPI_ISL_526713|2  ···················· ···················· ···················· ···················· ···················· ····················
```

| | | | | | | | | |
|---|---|---|---|---|---|---|---|---|
| G | 2750 | hCoV-19/South Korea/KCDC2780/2020\|EPI_ISL_526714\|2 | ···················· | ···················· | ···················· | ···················· | ···················· | ···················· |
| G | 2751 | hCoV-19/South Korea/KCDC2781/2020\|EPI_ISL_526715\|2 | ···················· | ···················· | ···················· | ···················· | ···················· | ···················· |
| G | 2752 | hCoV-19/South Korea/KCDC2782/2020\|EPI_ISL_526716\|2 | ···················· | ···················· | ···················· | ···················· | ···················· | ···················· |
| G | 2753 | hCoV-19/South Korea/KCDC2783/2020\|EPI_ISL_526717\|2 | ···················· | ···················· | ···················· | ···················· | ···················· | ···················· |
| G | 2754 | hCoV-19/South Korea/KCDC2784/2020\|EPI_ISL_526718\|2 | ···················· | ···················· | ···················· | ···················· | ···················· | ···················· |
| G | 2755 | hCoV-19/South Korea/KCDC2785/2020\|EPI_ISL_526719\|2 | ···················· | ···················· | ···················· | ···················· | ···················· | ···················· |
| G | 2756 | hCoV-19/South Korea/KCDC2786/2020\|EPI_ISL_526720\|2 | ···················· | ···················· | ···················· | ···················· | ···················· | ···················· |
| G | 2757 | hCoV-19/South Korea/KCDC2787/2020\|EPI_ISL_526721\|2 | ···················· | ···················· | ···················· | ···················· | ···················· | ···················· |
| G | 2758 | hCoV-19/South Korea/KCDC2788/2020\|EPI_ISL_526722\|2 | ···················· | ···················· | ···················· | ···················· | ···················· | ···················· |
| G | 2759 | hCoV-19/South Korea/KCDC2789/2020\|EPI_ISL_526723\|2 | ···················· | ···················· | ···················· | ···················· | ···················· | ···················· |
| G | 2760 | hCoV-19/South Korea/KCDC2790/2020\|EPI_ISL_526724\|2 | ···················· | ···················· | ···················· | ···················· | ···················· | ···················· |
| G | 2761 | hCoV-19/South Korea/KCDC2791/2020\|EPI_ISL_526725\|2 | ···················· | ···················· | ···················· | ···················· | ···················· | ···················· |
| G | 2762 | hCoV-19/South Korea/KCDC2792/2020\|EPI_ISL_526726\|2 | ···················· | ···················· | ···················· | ···················· | ···················· | ···················· |
| G | 2763 | hCoV-19/South Korea/KCDC2793/2020\|EPI_ISL_526727\|2 | ···················· | ···················· | ···················· | ···················· | ···················· | ···················· |
| G | 2764 | hCoV-19/South Korea/KCDC2794/2020\|EPI_ISL_526728\|2 | ···················· | ···················· | ···················· | ···················· | ···················· | ···················· |
| G | 2765 | hCoV-19/South Korea/KCDC2795/2020\|EPI_ISL_526729\|2 | ···················· | ···················· | ···················· | ···················· | ···················· | ···················· |
| G | 2766 | hCoV-19/South Korea/KCDC2796/2020\|EPI_ISL_526730\|2 | ···················· | ···················· | ···················· | ···················· | ···················· | ···················· |
| G | 2767 | hCoV-19/South Korea/KCDC2797/2020\|EPI_ISL_526731\|2 | ···················· | ···················· | ···················· | ···················· | ···················· | ···················· |
| G | 2768 | hCoV-19/South Korea/KCDC2798/2020\|EPI_ISL_526732\|2 | ···················· | ···················· | ···················· | ···················· | ···················· | ···················· |
| G | 2769 | hCoV-19/South Korea/KCDC2799/2020\|EPI_ISL_526733\|2 | ···················· | ···················· | ···················· | ···················· | ···················· | ···················· |
| G | 2770 | hCoV-19/South Korea/KCDC2800/2020\|EPI_ISL_526734\|2 | ···················· | ···················· | ···················· | ···················· | ···················· | ···················· |
| G | 2771 | hCoV-19/South Korea/KCDC2801/2020\|EPI_ISL_526735\|2 | ···················· | ···················· | ···················· | ···················· | ···················· | ···················· |
| G | 2772 | hCoV-19/South Korea/KCDC2802/2020\|EPI_ISL_526736\|2 | ···················· | ···················· | ···················· | ···················· | ···················· | ···················· |
| G | 2773 | hCoV-19/South Korea/KCDC2803/2020\|EPI_ISL_526737\|2 | ···················· | ···················· | ···················· | ···················· | ···················· | ···················· |
| G | 2774 | hCoV-19/South Korea/KCDC2804/2020\|EPI_ISL_526738\|2 | ···················· | ···················· | ···················· | ···················· | ···················· | ···················· |
| G | 2775 | hCoV-19/South Korea/KCDC2805/2020\|EPI_ISL_526739\|2 | ···················· | ···················· | ···················· | ···················· | ···················· | ···················· |
| G | 2776 | hCoV-19/South Korea/KCDC2806/2020\|EPI_ISL_526740\|2 | ···················· | ···················· | ···················· | ···················· | ···················· | ···················· |
| G | 2777 | hCoV-19/South Korea/KCDC2807/2020\|EPI_ISL_526741\|2 | ···················· | ···················· | ···················· | ···················· | ···················· | ···················· |
| G | 2778 | hCoV-19/South Korea/KCDC2808/2020\|EPI_ISL_526742\|2 | ···················· | ···················· | ···················· | ···················· | ···················· | ···················· |
| G | 2779 | hCoV-19/South Korea/KCDC2809/2020\|EPI_ISL_526743\|2 | ···················· | ···················· | ···················· | ···················· | ···················· | ···················· |
| G | 2780 | hCoV-19/South Korea/KCDC2810/2020\|EPI_ISL_526744\|2 | ···················· | ···················· | ···················· | ···················· | ···················· | ···················· |
| G | 2781 | hCoV-19/South Korea/KCDC2811/2020\|EPI_ISL_526745\|2 | ···················· | ···················· | ···················· | ···················· | ···················· | ···················· |
| G | 2782 | hCoV-19/South Korea/KCDC2813/2020\|EPI_ISL_526746\|2 | ···················· | ···················· | ···················· | ···················· | ···················· | ···················· |
| G | 2783 | hCoV-19/England/201690346/2020\|EPI_ISL_526754\|2020 | ···················· | ···················· | ···················· | ···················· | ···················· | ···················· |
| G | 2784 | hCoV-19/England/201720576/2020\|EPI_ISL_526755\|2020 | ···················· | ···················· | ···················· | ···················· | ···················· | ···················· |
| G | 2785 | hCoV-19/England/201720577/2020\|EPI_ISL_526756\|2020 | ···················· | ···················· | ···················· | ···················· | ···················· | ···················· |
| G | 2786 | hCoV-19/England/201780026/2020\|EPI_ISL_526757\|2020 | ···················· | ···················· | ···················· | ···················· | ···················· | ···················· |
| G | 2787 | hCoV-19/England/201780590/2020\|EPI_ISL_526758\|2020 | ···················· | ···················· | ···················· | ···················· | ···················· | ···················· |
| G | 2788 | hCoV-19/England/201780626/2020\|EPI_ISL_526759\|2020 | ···················· | ···················· | ···················· | ···················· | ···················· | ···················· |
| G | 2789 | hCoV-19/England/201790453/2020\|EPI_ISL_526762\|2020 | ···················· | ···················· | ···················· | ···················· | ···················· | ···················· |
| G | 2790 | hCoV-19/England/201790455/2020\|EPI_ISL_526763\|2020 | ···················· | ···················· | ···················· | ···················· | ···················· | ···················· |
| G | 2791 | hCoV-19/England/201790456/2020\|EPI_ISL_526764\|2020 | ···················· | ···················· | ···················· | ···················· | ···················· | ···················· |
| G | 2792 | hCoV-19/England/201790459/2020\|EPI_ISL_526765\|2020 | ···················· | ···················· | ···················· | ···················· | ···················· | ···················· |
| G | 2793 | hCoV-19/England/201790472/2020\|EPI_ISL_526769\|2020 | ···················· | ···················· | ···················· | ···················· | ···················· | ···················· |
| G | 2794 | hCoV-19/USA/VA-DCLS-0483/2020\|EPI_ISL_526790\|2020- | ···················· | ···················· | ···················· | ···················· | ···················· | ···················· |
| G | 2795 | hCoV-19/USA/VA-DCLS-0484/2020\|EPI_ISL_526791\|2020- | ···················· | ···················· | ···················· | ···················· | ···················· | ···················· |
| G | 2796 | hCoV-19/USA/VA-DCLS-0485/2020\|EPI_ISL_526792\|2020- | ···················· | ···················· | ···················· | ···················· | ···················· | ···················· |
| G | 2797 | hCoV-19/USA/VA-DCLS-0486/2020\|EPI_ISL_526793\|2020- | ···················· | ···················· | ···················· | ···················· | ···················· | ···················· |
| G | 2798 | hCoV-19/USA/VA-DCLS-0487/2020\|EPI_ISL_526794\|2020- | ···················· | ···················· | ···················· | ···················· | ···················· | ···················· |
| G | 2799 | hCoV-19/USA/VA-DCLS-0488/2020\|EPI_ISL_526795\|2020- | ···················· | ···················· | ···················· | ···················· | ···················· | ···················· |
| G | 2800 | hCoV-19/USA/VA-DCLS-0489/2020\|EPI_ISL_526796\|2020- | ···················· | ···················· | ···················· | ···················· | ···················· | ········A··········· |
| G | 2801 | hCoV-19/USA/VA-DCLS-0575/2020\|EPI_ISL_526797\|2020- | ···················· | ···················· | ···················· | ···················· | ···················· | ···················· |
| G | 2802 | hCoV-19/USA/VA-DCLS-0577/2020\|EPI_ISL_526798\|2020- | ···················· | ···················· | ···················· | ···················· | ···················· | ···················· |
| G | 2803 | hCoV-19/USA/VA-DCLS-0578/2020\|EPI_ISL_526799\|2020- | ···················· | ···················· | ···················· | ···················· | ···················· | ···················· |
| G | 2804 | hCoV-19/Italy/ABR-IZSGC-TE92551/2020\|EPI_ISL_52901 | ···················· | ···················· | ···················· | ···················· | ···················· | ···················· |
| G | 2805 | hCoV-19/Italy/ABR-IZSGC-TE92552/2020\|EPI_ISL_52901 | ···················· | ···················· | ···················· | ···················· | ···················· | ···················· |
| G | 2806 | hCoV-19/Italy/ABR-IZSGC-TE93935/2020\|EPI_ISL_52901 | ···················· | ···················· | ···················· | ···················· | ···················· | ···················· |
| G | 2807 | hCoV-19/Italy/ABR-IZSGC-TE94745/2020\|EPI_ISL_52901 | ···················· | ···················· | ···················· | ···················· | ···················· | ···················· |
| G | 2808 | hCoV-19/Italy/ABR-IZSGC-TE99752/2020\|EPI_ISL_52901 | ···················· | ···················· | ···················· | ···················· | ···················· | ···················· |
| G | 2809 | hCoV-19/Italy/ABR-IZSGC-TE99753/2020\|EPI_ISL_52901 | ···················· | ···················· | ···················· | ···················· | ···················· | ···················· |
| G | 2810 | hCoV-19/Italy/ABR-IZSGC-TE10151/2020\|EPI_ISL_52901 | ···················· | ···················· | ···················· | ···················· | ···················· | ···················· |
| G | 2811 | hCoV-19/Italy/ABR-IZSGC-TE18513/2020\|EPI_ISL_52901 | ···················· | ···················· | ···················· | ···················· | ···················· | ···················· |
| G | 2812 | hCoV-19/Italy/ABR-IZSGC-TE27016/2020\|EPI_ISL_52901 | ···················· | ···················· | ···················· | ···················· | ···················· | ···················· |
| G | 2813 | hCoV-19/Spain/MD-H12-69/2020\|EPI_ISL_530000\|2020-0 | ···················· | ···················· | ···················· | ···················· | ···················· | ···················· |
| G | 2814 | hCoV-19/Spain/MD-H12-70/2020\|EPI_ISL_530001\|2020-0 | ···················· | ···················· | ···················· | ···················· | ···················· | ···················· |
| G | 2815 | hCoV-19/Spain/MD-H12-71/2020\|EPI_ISL_530002\|2020-0 | ···················· | ···················· | ···················· | ···················· | ···················· | ···················· |
| G | 2816 | hCoV-19/Spain/MD-H12-72/2020\|EPI_ISL_530003\|2020-0 | ···················· | ···················· | ···················· | ···················· | ···················· | ···················· |
| G | 2817 | hCoV-19/England/CAMB-1B5BAE/2020\|EPI_ISL_517000\|20 | ···················· | ···················· | ···················· | ···················· | ···················· | ···················· |
| G | 2818 | hCoV-19/England/LIVE-A72F6/2020\|EPI_ISL_517001\|202 | ···················· | ···················· | ···················· | ···················· | ···················· | ···················· |
| G | 2821 | hCoV-19/England/LIVE-A7320/2020\|EPI_ISL_517004\|202 | ···················· | ·A·················· | ···················· | ···················· | ···················· | ···················· |
| G | 2825 | hCoV-19/England/LIVE-A736C/2020\|EPI_ISL_517008\|202 | ···················· | ···················· | ···················· | ···················· | ···················· | ···················· |
| G | 2827 | hCoV-19/England/LIVE-A7399/2020\|EPI_ISL_517011\|202 | ···················· | ···················· | ···················· | ···················· | ···················· | ···················· |
| G | 2828 | hCoV-19/England/LIVE-A73A8/2020\|EPI_ISL_517012\|202 | ···················· | ···················· | ···················· | ···················· | ···················· | ···················· |
| G | 2831 | hCoV-19/England/LIVE-A73D5/2020\|EPI_ISL_517015\|202 | ···················· | ···················· | ···················· | ···················· | ···················· | ···················· |
| G | 2835 | hCoV-19/England/LIVE-A742D/2020\|EPI_ISL_517020\|202 | ···················· | ···················· | ···················· | ···················· | ···················· | ···················· |
| G | 2836 | hCoV-19/England/LIVE-A743C/2020\|EPI_ISL_517021\|202 | ···················· | ···················· | ···················· | ···················· | ···················· | ···················· |
| G | 2839 | hCoV-19/England/LIVE-A7469/2020\|EPI_ISL_517024\|202 | ···················· | ···················· | ···················· | ···················· | ···················· | ···················· |
| G | 2840 | hCoV-19/England/LIVE-A7478/2020\|EPI_ISL_517025\|202 | ···················· | ···················· | ···················· | ···················· | ···················· | ···················· |
| G | 2844 | hCoV-19/Australia/VIC6312/2020\|EPI_ISL_518300\|2020 | ···················· | ···················· | ···················· | ···················· | ···················· | ···················· |
| G | 2845 | hCoV-19/Australia/VIC6313/2020\|EPI_ISL_518301\|2020 | ···················· | ···················· | ···················· | ···················· | ···················· | ···················· |

| | | | | | | | | |
|---|---|---|---|---|---|---|---|---|
| G | 2846 | hCoV-19/Australia/VIC6314/2020\|EPI_ISL_518302\|2020 | .................... | .................... | .................... | .................... | .................... | .................... |
| G | 2847 | hCoV-19/Australia/VIC6316/2020\|EPI_ISL_518304\|2020 | .................... | .................... | .................... | .................... | .................... | .................... |
| G | 2848 | hCoV-19/Australia/VIC6317/2020\|EPI_ISL_518305\|2020 | .................... | .................... | .................... | .................... | .................... | .................... |
| G | 2849 | hCoV-19/Australia/VIC6318/2020\|EPI_ISL_518306\|2020 | .................... | .................... | .................... | .................... | .................... | .................... |
| G | 2850 | hCoV-19/Australia/VIC6319/2020\|EPI_ISL_518307\|2020 | .................... | .................... | .................... | .................... | .................... | .................... |
| G | 2851 | hCoV-19/Australia/VIC6320/2020\|EPI_ISL_518308\|2020 | .................... | .................... | .................... | .................... | .................... | .................... |
| G | 2852 | hCoV-19/Australia/VIC6322/2020\|EPI_ISL_518309\|2020 | .................... | .................... | .................... | .................... | .................... | .................... |
| G | 2853 | hCoV-19/Australia/VIC6324/2020\|EPI_ISL_518311\|2020 | .................... | .................... | .................... | .................... | .................... | .................... |
| G | 2854 | hCoV-19/Australia/VIC6325/2020\|EPI_ISL_518312\|2020 | .................... | .................... | .................... | .................... | .................... | .................... |
| G | 2855 | hCoV-19/Australia/VIC6326/2020\|EPI_ISL_518313\|2020 | .................... | .................... | .................... | .................... | .................... | .................... |
| G | 2856 | hCoV-19/Australia/VIC6327/2020\|EPI_ISL_518314\|2020 | .................... | .................... | .................... | .................... | .................... | .................... |
| G | 2857 | hCoV-19/Australia/VIC6332/2020\|EPI_ISL_518316\|2020 | .................... | .................... | .................... | .................... | .................... | .................... |
| G | 2858 | hCoV-19/Australia/VIC6335/2020\|EPI_ISL_518318\|2020 | .................... | .................... | .................... | .................... | .................... | .................... |
| G | 2859 | hCoV-19/Australia/VIC6336/2020\|EPI_ISL_518319\|2020 | .................... | .................... | .................... | .................... | .................... | .................... |
| G | 2860 | hCoV-19/Australia/VIC6339/2020\|EPI_ISL_518321\|2020 | .................... | .................... | .................... | .................... | .................... | .................... |
| G | 2861 | hCoV-19/Australia/VIC6340/2020\|EPI_ISL_518322\|2020 | .................... | .................... | .................... | .................... | .................... | .................... |
| G | 2862 | hCoV-19/Australia/VIC6341/2020\|EPI_ISL_518323\|2020 | .................... | .................... | .................... | .................... | .................... | .................... |
| G | 2863 | hCoV-19/Australia/VIC6342/2020\|EPI_ISL_518324\|2020 | .................... | .................... | .................... | .................... | .................... | .................... |
| G | 2864 | hCoV-19/Australia/VIC6343/2020\|EPI_ISL_518325\|2020 | .................... | .................... | .................... | .................... | .................... | .................... |
| G | 2865 | hCoV-19/Australia/VIC6346/2020\|EPI_ISL_518326\|2020 | .................... | .................... | .................... | .................... | .................... | .................... |
| G | 2866 | hCoV-19/Australia/VIC6348/2020\|EPI_ISL_518328\|2020 | .................... | .................... | .................... | .................... | .................... | .................... |
| G | 2867 | hCoV-19/Australia/VIC6349/2020\|EPI_ISL_518329\|2020 | .................... | .................... | .................... | .................... | .................... | .................... |
| G | 2868 | hCoV-19/Australia/VIC6350/2020\|EPI_ISL_518330\|2020 | .................... | .................... | .................... | .................... | .................... | .................... |
| G | 2869 | hCoV-19/Australia/VIC6351/2020\|EPI_ISL_518331\|2020 | .................... | .................... | .................... | .................... | .................... | .................... |
| G | 2870 | hCoV-19/Australia/VIC6352/2020\|EPI_ISL_518332\|2020 | .................... | .................... | .................... | .................... | .................... | .................... |
| G | 2871 | hCoV-19/Australia/VIC6353/2020\|EPI_ISL_518333\|2020 | .................... | .................... | .................... | .................... | .................... | .................... |
| G | 2872 | hCoV-19/Australia/VIC6354/2020\|EPI_ISL_518334\|2020 | .................... | .................... | .................... | .................... | .................... | .................... |
| G | 2873 | hCoV-19/Australia/VIC6355/2020\|EPI_ISL_518335\|2020 | .................... | .................... | .................... | .................... | .................... | .................... |
| G | 2874 | hCoV-19/Australia/VIC6356/2020\|EPI_ISL_518336\|2020 | .................... | .................... | .................... | .................... | .................... | .................... |
| G | 2875 | hCoV-19/Australia/VIC6357/2020\|EPI_ISL_518337\|2020 | .................... | .................... | .................... | .................... | .................... | .................... |
| G | 2876 | hCoV-19/Australia/VIC5503/2020\|EPI_ISL_518339\|2020 | .................... | .................... | .................... | .................... | .................... | .................... |
| G | 2877 | hCoV-19/Australia/VIC5504/2020\|EPI_ISL_518340\|2020 | .................... | .................... | .................... | .................... | .................... | .................... |
| G | 2878 | hCoV-19/Australia/VIC5505/2020\|EPI_ISL_518341\|2020 | .................... | .................... | .................... | .................... | .................... | .................... |
| G | 2879 | hCoV-19/Australia/VIC5506/2020\|EPI_ISL_518342\|2020 | .................... | .................... | .................... | .................... | .................... | .................... |
| G | 2880 | hCoV-19/Australia/VIC5507/2020\|EPI_ISL_518343\|2020 | .................... | .................... | .................... | .................... | .................... | .................... |
| G | 2881 | hCoV-19/Australia/VIC5508/2020\|EPI_ISL_518344\|2020 | .................... | .................... | .................... | .................... | .................... | .................... |
| G | 2882 | hCoV-19/Australia/VIC5509/2020\|EPI_ISL_518345\|2020 | .................... | .................... | .................... | .................... | .................... | .................... |
| G | 2883 | hCoV-19/Australia/VIC5510/2020\|EPI_ISL_518346\|2020 | .................... | .................... | .................... | .................... | .................... | .................... |
| G | 2884 | hCoV-19/Australia/VIC5512/2020\|EPI_ISL_518347\|2020 | .................... | .................... | .................... | .................... | .................... | .................... |
| G | 2885 | hCoV-19/Australia/VIC5514/2020\|EPI_ISL_518349\|2020 | .................... | .................... | .................... | .................... | .................... | .................... |
| G | 2886 | hCoV-19/Australia/VIC5515/2020\|EPI_ISL_518350\|2020 | .................... | .................... | .................... | .................... | .................... | .................... |
| G | 2887 | hCoV-19/Australia/VIC5516/2020\|EPI_ISL_518351\|2020 | .................... | .................... | .................... | .................... | .................... | .................... |
| G | 2888 | hCoV-19/Australia/VIC5517/2020\|EPI_ISL_518352\|2020 | .................... | .................... | .................... | .................... | .................... | .................... |
| G | 2889 | hCoV-19/Australia/VIC5518/2020\|EPI_ISL_518353\|2020 | .................... | .................... | .................... | .................... | .................... | .................... |
| G | 2890 | hCoV-19/Australia/VIC5519/2020\|EPI_ISL_518354\|2020 | .................... | .................... | .................... | .................... | .................... | .................... |
| G | 2891 | hCoV-19/Australia/VIC5521/2020\|EPI_ISL_518355\|2020 | .................... | .................... | .................... | .................... | .................... | .................... |
| G | 2892 | hCoV-19/Australia/VIC5522/2020\|EPI_ISL_518356\|2020 | .................... | .................... | .................... | .................... | .................... | .................... |
| G | 2893 | hCoV-19/Australia/VIC5523/2020\|EPI_ISL_518357\|2020 | .................... | .................... | .................... | .................... | .................... | .................... |
| G | 2894 | hCoV-19/Australia/VIC5524/2020\|EPI_ISL_518358\|2020 | .................... | .................... | .................... | .................... | .................... | .................... |
| G | 2895 | hCoV-19/Australia/VIC5525/2020\|EPI_ISL_518359\|2020 | .................... | .................... | .................... | .................... | .................... | .................... |
| G | 2896 | hCoV-19/Australia/VIC5538/2020\|EPI_ISL_518369\|2020 | .................... | .................... | .................... | .................... | .................... | .................... |
| G | 2897 | hCoV-19/Australia/VIC4335/2020\|EPI_ISL_519631\|2020 | .................... | .................... | .................... | .................... | .................... | .................... |
| G | 2898 | hCoV-19/Australia/VIC4336/2020\|EPI_ISL_519632\|2020 | .................... | .................... | .................... | .................... | .................... | .................... |
| G | 2899 | hCoV-19/Australia/VIC4338/2020\|EPI_ISL_519634\|2020 | .................... | .................... | .................... | .................... | .................... | .................... |
| G | 2900 | hCoV-19/Australia/VIC4339/2020\|EPI_ISL_519635\|2020 | .................... | .................... | .................... | .................... | .................... | .................... |
| G | 2901 | hCoV-19/Australia/VIC4340/2020\|EPI_ISL_519636\|2020 | .................... | .................... | .................... | .................... | .................... | .................... |
| G | 2902 | hCoV-19/Australia/VIC4341/2020\|EPI_ISL_519637\|2020 | .................... | .................... | .................... | .................... | .................... | .................... |
| G | 2903 | hCoV-19/Australia/VIC4342/2020\|EPI_ISL_519638\|2020 | .................... | .................... | .................... | .................... | .................... | .................... |
| G | 2904 | hCoV-19/Australia/VIC4343/2020\|EPI_ISL_519639\|2020 | .................... | .................... | .................... | .................... | .................... | .................... |
| G | 2905 | hCoV-19/Australia/VIC4344/2020\|EPI_ISL_519640\|2020 | .................... | .................... | .................... | .................... | .................... | .................... |
| G | 2906 | hCoV-19/Australia/VIC4345/2020\|EPI_ISL_519641\|2020 | .................... | .................... | .................... | .................... | .................... | .................... |
| G | 2907 | hCoV-19/Australia/VIC4346/2020\|EPI_ISL_519642\|2020 | .................... | .................... | .................... | .................... | .................... | .................... |
| G | 2908 | hCoV-19/Australia/VIC4347/2020\|EPI_ISL_519643\|2020 | .................... | .................... | .................... | .................... | .................... | .................... |
| G | 2909 | hCoV-19/Australia/VIC4349/2020\|EPI_ISL_519645\|2020 | .................... | .................... | .................... | .................... | .................... | .................... |
| G | 2910 | hCoV-19/Australia/VIC4350/2020\|EPI_ISL_519646\|2020 | .................... | .................... | .................... | .................... | .................... | .................... |
| G | 2911 | hCoV-19/Australia/VIC4351/2020\|EPI_ISL_519647\|2020 | .................... | .................... | .................... | .................... | .................... | .................... |
| G | 2912 | hCoV-19/Australia/VIC4352/2020\|EPI_ISL_519648\|2020 | .................... | .................... | .................... | .................... | .................... | .................... |
| G | 2913 | hCoV-19/Australia/VIC4353/2020\|EPI_ISL_519649\|2020 | .................... | .................... | .................... | .................... | .................... | .................... |
| G | 2914 | hCoV-19/Australia/VIC4354/2020\|EPI_ISL_519650\|2020 | .................... | .................... | .................... | .................... | .................... | .................... |
| G | 2915 | hCoV-19/Australia/VIC4355/2020\|EPI_ISL_519651\|2020 | .................... | .................... | .................... | .................... | .................... | .................... |
| G | 2916 | hCoV-19/Australia/VIC4356/2020\|EPI_ISL_519652\|2020 | .................... | .................... | .................... | .................... | .................... | .................... |
| G | 2917 | hCoV-19/Australia/VIC4357/2020\|EPI_ISL_519653\|2020 | .................... | .................... | .................... | .................... | .................... | .................... |
| G | 2918 | hCoV-19/Australia/VIC4358/2020\|EPI_ISL_519654\|2020 | .................... | .................... | .................... | .................... | .................... | .................... |
| G | 2919 | hCoV-19/Australia/VIC4359/2020\|EPI_ISL_519655\|2020 | .................... | .................... | .................... | .................... | .................... | .................... |
| G | 2920 | hCoV-19/Australia/VIC4360/2020\|EPI_ISL_519656\|2020 | .................... | .................... | .................... | .................... | .................... | .................... |
| G | 2921 | hCoV-19/Australia/VIC4361/2020\|EPI_ISL_519657\|2020 | .................... | .................... | .................... | .................... | .................... | .................... |
| G | 2922 | hCoV-19/Australia/VIC4362/2020\|EPI_ISL_519658\|2020 | .................... | .................... | .................... | .................... | .................... | .................... |
| G | 2923 | hCoV-19/Australia/VIC4363/2020\|EPI_ISL_519659\|2020 | .................... | .................... | .................... | .................... | .................... | .................... |
| G | 2924 | hCoV-19/Australia/VIC4364/2020\|EPI_ISL_519660\|2020 | .................... | .................... | .................... | .................... | .................... | .................... |
| G | 2925 | hCoV-19/Australia/VIC4365/2020\|EPI_ISL_519661\|2020 | .................... | .................... | .................... | .................... | .................... | .................... |

| | | | | | | | | |
|---|---|---|---|---|---|---|---|---|
| G | 2926 hCoV-19/Australia/VIC4366/2020\|EPI_ISL_519662\|2020 | ···················· | ···················· | ····················· | ···················· | ···················· | ····················· | |
| G | 2927 hCoV-19/Australia/VIC4367/2020\|EPI_ISL_519663\|2020 | ···················· | ···················· | ····················· | ···················· | ···················· | ····················· | |
| G | 2928 hCoV-19/Australia/VIC4368/2020\|EPI_ISL_519664\|2020 | ···················· | ···················· | ····················· | ···················· | ···················· | ····················· | |
| G | 2929 hCoV-19/Australia/VIC4369/2020\|EPI_ISL_519665\|2020 | ···················· | ···················· | ····················· | ···················· | ···················· | ····················· | |
| G | 2930 hCoV-19/Australia/VIC4370/2020\|EPI_ISL_519666\|2020 | ···················· | ···················· | ····················· | ···················· | ···················· | ····················· | |
| G | 2931 hCoV-19/Australia/VIC4371/2020\|EPI_ISL_519667\|2020 | ···················· | ···················· | ····················· | ···················· | ···················· | ····················· | |
| G | 2932 hCoV-19/Australia/VIC4372/2020\|EPI_ISL_519668\|2020 | ···················· | ···················· | ····················· | ···················· | ···················· | ····················· | |
| G | 2933 hCoV-19/Australia/VIC4373/2020\|EPI_ISL_519669\|2020 | ···················· | ···················· | ····················· | ···················· | ···················· | ····················· | |
| G | 2934 hCoV-19/Australia/VIC4374/2020\|EPI_ISL_519670\|2020 | ···················· | ···················· | ····················· | ···················· | ···················· | ····················· | |
| G | 2935 hCoV-19/Australia/VIC4375/2020\|EPI_ISL_519671\|2020 | ···················· | ···················· | ····················· | ···················· | ···················· | ····················· | |
| G | 2936 hCoV-19/Australia/VIC4376/2020\|EPI_ISL_519672\|2020 | ···················· | ···················· | ····················· | ···················· | ···················· | ····················· | |
| G | 2937 hCoV-19/Australia/VIC4378/2020\|EPI_ISL_519674\|2020 | ···················· | ···················· | ····················· | ···················· | ···················· | ····················· | |
| G | 2938 hCoV-19/Australia/VIC4379/2020\|EPI_ISL_519675\|2020 | ···················· | ···················· | ····················· | ···················· | ···················· | ····················· | |
| G | 2939 hCoV-19/Australia/VIC4380/2020\|EPI_ISL_519676\|2020 | ···················· | ···················· | ····················· | ···················· | ···················· | ····················· | |
| G | 2940 hCoV-19/Australia/VIC4381/2020\|EPI_ISL_519677\|2020 | ···················· | ···················· | ····················· | ···················· | ···················· | ····················· | |
| G | 2941 hCoV-19/Australia/VIC4382/2020\|EPI_ISL_519678\|2020 | ···················· | ···················· | ····················· | ···················· | ···················· | ····················· | |
| G | 2942 hCoV-19/Australia/VIC4383/2020\|EPI_ISL_519679\|2020 | ···················· | ···················· | ····················· | ···················· | ···················· | ····················· | |
| G | 2943 hCoV-19/Australia/VIC4384/2020\|EPI_ISL_519680\|2020 | ···················· | ···················· | ····················· | ···················· | ···················· | ····················· | |
| G | 2944 hCoV-19/Australia/VIC4385/2020\|EPI_ISL_519681\|2020 | ···················· | ···················· | ····················· | ···················· | ···················· | ····················· | |
| G | 2945 hCoV-19/Australia/VIC4386/2020\|EPI_ISL_519682\|2020 | ···················· | ···················· | ····················· | ···················· | ···················· | ····················· | |
| G | 2946 hCoV-19/Australia/VIC4387/2020\|EPI_ISL_519683\|2020 | ···················· | ···················· | ····················· | ···················· | ···················· | ····················· | |
| G | 2947 hCoV-19/Australia/VIC4388/2020\|EPI_ISL_519684\|2020 | ···················· | ···················· | ····················· | ···················· | ···················· | ····················· | |
| G | 2948 hCoV-19/Australia/VIC4389/2020\|EPI_ISL_519685\|2020 | ···················· | ···················· | ····················· | ···················· | ···················· | ····················· | |
| G | 2949 hCoV-19/Australia/VIC4390/2020\|EPI_ISL_519686\|2020 | ···················· | ···················· | ····················· | ···················· | ···················· | ····················· | |
| G | 2950 hCoV-19/Australia/VIC4391/2020\|EPI_ISL_519687\|2020 | ···················· | ···················· | ····················· | ···················· | ···················· | ····················· | |
| G | 2951 hCoV-19/Australia/VIC4392/2020\|EPI_ISL_519688\|2020 | ···················· | ···················· | ····················· | ···················· | ···················· | ····················· | |
| G | 2952 hCoV-19/Australia/VIC4393/2020\|EPI_ISL_519689\|2020 | ···················· | ···················· | ····················· | ···················· | ···················· | ····················· | |
| G | 2953 hCoV-19/Australia/VIC4403/2020\|EPI_ISL_519699\|2020 | ···················· | ···················· | ····················· | ···················· | ···················· | ····················· | |
| G | 2954 hCoV-19/Australia/VIC3898/2020\|EPI_ISL_520610\|2020 | ···················· | ···················· | ····················· | ···················· | ···················· | ····················· | |
| G | 2955 hCoV-19/Australia/VIC3899/2020\|EPI_ISL_520611\|2020 | ···················· | ···················· | ····················· | ···················· | ···················· | ····················· | |
| G | 2956 hCoV-19/Australia/VIC3900/2020\|EPI_ISL_520612\|2020 | ···················· | ···················· | ····················· | ···················· | ···················· | ····················· | |
| G | 2957 hCoV-19/Australia/VIC3901/2020\|EPI_ISL_520613\|2020 | ···················· | ···················· | ····················· | ···················· | ···················· | ····················· | |
| G | 2958 hCoV-19/Australia/VIC3902/2020\|EPI_ISL_520614\|2020 | ···················· | ···················· | ····················· | ···················· | ···················· | ····················· | |
| G | 2959 hCoV-19/Australia/VIC3903/2020\|EPI_ISL_520615\|2020 | ···················· | ···················· | ····················· | ···················· | ···················· | ····················· | |
| G | 2960 hCoV-19/Australia/VIC3904/2020\|EPI_ISL_520616\|2020 | ···················· | ···················· | ····················· | ···················· | ···················· | ····················· | |
| G | 2961 hCoV-19/Australia/VIC3905/2020\|EPI_ISL_520617\|2020 | ···················· | ···················· | ····················· | ···················· | ···················· | ····················· | |
| G | 2962 hCoV-19/Australia/VIC3906/2020\|EPI_ISL_520618\|2020 | ···················· | ···················· | ····················· | ···················· | ···················· | ····················· | |
| G | 2963 hCoV-19/Australia/VIC3907/2020\|EPI_ISL_520619\|2020 | ···················· | ···················· | ····················· | ···················· | ···················· | ····················· | |
| G | 2964 hCoV-19/Australia/VIC3908/2020\|EPI_ISL_520620\|2020 | ···················· | ···················· | ····················· | ···················· | ···················· | ····················· | |
| G | 2965 hCoV-19/Australia/VIC3909/2020\|EPI_ISL_520621\|2020 | ···················· | ···················· | ····················· | ···················· | NNN················· | ····················· | 1 |
| G | 2966 hCoV-19/Australia/VIC3910/2020\|EPI_ISL_520622\|2020 | ···················· | ···················· | ····················· | ···················· | ···················· | ····················· | |
| G | 2967 hCoV-19/Australia/VIC3911/2020\|EPI_ISL_520623\|2020 | ···················· | ···················· | ····················· | ···················· | ···················· | ····················· | |
| G | 2968 hCoV-19/Australia/VIC3912/2020\|EPI_ISL_520624\|2020 | ···················· | ···················· | ····················· | ···················· | ···················· | ····················· | |
| G | 2969 hCoV-19/Australia/VIC3913/2020\|EPI_ISL_520625\|2020 | ···················· | ···················· | ····················· | ···················· | ···················· | ····················· | |
| G | 2970 hCoV-19/Australia/VIC3914/2020\|EPI_ISL_520626\|2020 | ···················· | ···················· | ····················· | ···················· | ···················· | ····················· | |
| G | 2971 hCoV-19/Australia/VIC3915/2020\|EPI_ISL_520627\|2020 | ···················· | ···················· | ····················· | ···················· | ···················· | ····················· | |
| G | 2972 hCoV-19/Australia/VIC3916/2020\|EPI_ISL_520628\|2020 | ···················· | ···················· | ····················· | ···················· | ···················· | ····················· | |
| G | 2973 hCoV-19/Australia/VIC3917/2020\|EPI_ISL_520629\|2020 | ···················· | ···················· | ····················· | ···················· | ···················· | ····················· | |
| G | 2974 hCoV-19/Australia/VIC3918/2020\|EPI_ISL_520630\|2020 | ···················· | ···················· | ····················· | ···················· | ···················· | ····················· | |
| G | 2975 hCoV-19/Australia/VIC3919/2020\|EPI_ISL_520631\|2020 | ···················· | ···················· | ····················· | ···················· | ···················· | ····················· | |
| G | 2976 hCoV-19/Australia/VIC3920/2020\|EPI_ISL_520632\|2020 | ···················· | ···················· | ····················· | ···················· | ···················· | ····················· | |
| G | 2977 hCoV-19/Australia/VIC3921/2020\|EPI_ISL_520633\|2020 | ···················· | ···················· | ····················· | ···················· | ···················· | ····················· | |
| G | 2978 hCoV-19/Australia/VIC3922/2020\|EPI_ISL_520634\|2020 | ···················· | ···················· | ····················· | ···················· | ···················· | ····················· | |
| G | 2980 hCoV-19/Australia/VIC3924/2020\|EPI_ISL_520636\|2020 | ···················· | ···················· | ····················· | ···················· | ···················· | ····················· | |
| G | 2981 hCoV-19/Australia/VIC3925/2020\|EPI_ISL_520637\|2020 | ···················· | ···················· | ····················· | ···················· | ···················· | ····················· | |
| G | 2982 hCoV-19/Australia/VIC3926/2020\|EPI_ISL_520638\|2020 | ···················· | ···················· | ····················· | ···················· | ···················· | ····················· | |
| G | 2983 hCoV-19/Australia/VIC3927/2020\|EPI_ISL_520639\|2020 | ···················· | ···················· | ····················· | ···················· | ···················· | ····················· | |
| G | 2984 hCoV-19/Australia/VIC3928/2020\|EPI_ISL_520640\|2020 | ···················· | ···················· | ····················· | ···················· | ···················· | ····················· | |
| G | 2985 hCoV-19/Australia/VIC3929/2020\|EPI_ISL_520641\|2020 | ···················· | ···················· | ····················· | ···················· | ·NNN················ | ····················· | 1 |
| G | 2986 hCoV-19/Australia/VIC3930/2020\|EPI_ISL_520642\|2020 | ···················· | ···················· | ····················· | ···················· | ···················· | ····················· | |
| G | 2987 hCoV-19/Australia/VIC3931/2020\|EPI_ISL_520643\|2020 | ···················· | ···················· | ····················· | ···················· | ···················· | ····················· | |
| G | 2989 hCoV-19/Australia/VIC3934/2020\|EPI_ISL_520646\|2020 | ···················· | ···················· | ····················· | ···················· | ···················· | ····················· | |
| G | 2990 hCoV-19/Australia/VIC3935/2020\|EPI_ISL_520647\|2020 | ···················· | ···················· | ····················· | ···················· | NNN················· | ····················· | 1 |
| G | 2991 hCoV-19/Australia/VIC3936/2020\|EPI_ISL_520648\|2020 | ···················· | ···················· | ····················· | ···················· | ···················· | ····················· | |
| G | 2992 hCoV-19/Australia/VIC3938/2020\|EPI_ISL_520649\|2020 | ···················· | ···················· | ····················· | ···················· | ···················· | ····················· | |
| G | 2994 hCoV-19/Australia/VIC3940/2020\|EPI_ISL_520651\|2020 | ···················· | ···················· | ····················· | ···················· | ···················· | ····················· | |
| G | 2995 hCoV-19/Australia/VIC3941/2020\|EPI_ISL_520652\|2020 | ···················· | ···················· | ····················· | ···················· | ···················· | ····················· | |
| G | 2996 hCoV-19/Australia/VIC3942/2020\|EPI_ISL_520653\|2020 | ···················· | ···················· | ····················· | ···················· | ···················· | ····················· | |
| G | 2998 hCoV-19/Australia/VIC3945/2020\|EPI_ISL_520656\|2020 | ···················· | ···················· | ····················· | ···················· | ···················· | ····················· | |
| G | 2999 hCoV-19/Australia/VIC3946/2020\|EPI_ISL_520657\|2020 | ···················· | ···················· | ····················· | ···················· | ···················· | ····················· | |
| G | 3000 hCoV-19/Australia/VIC3947/2020\|EPI_ISL_520658\|2020 | ···················· | ···················· | ····················· | ···················· | ···················· | ····················· | |
| G | 3001 hCoV-19/Australia/VIC3948/2020\|EPI_ISL_520659\|2020 | ···················· | ···················· | ····················· | ···················· | ···················· | ····················· | |
| G | 3003 hCoV-19/Australia/VIC3950/2020\|EPI_ISL_520661\|2020 | ···················· | ···················· | ····················· | ···················· | ···················· | ····················· | |
| G | 3004 hCoV-19/Australia/VIC3951/2020\|EPI_ISL_520662\|2020 | ···················· | ···················· | ····················· | ···················· | ···················· | ····················· | |
| G | 3005 hCoV-19/United Arab Emirates/L104447095/2020\|EPI_I | ···················· | ···················· | ····················· | ···················· | ···················· | ····················· | |
| G | 3006 hCoV-19/United Arab Emirates/L109275764/2020\|EPI_I | ···················· | ···················· | ····················· | ···················· | ···················· | ····················· | |
| G | 3007 hCoV-19/United Arab Emirates/L121759318/2020\|EPI_I | ···················· | ···················· | ····················· | ···················· | ···················· | ····················· | |
| G | 3008 hCoV-19/United Arab Emirates/L123065607/2020\|EPI_I | ···················· | ···················· | ····················· | ···················· | ···················· | ····················· | |
| G | 3009 hCoV-19/United Arab Emirates/L2004034430/2020\|EPI_ | ···················· | ···················· | ····················· | ···················· | ···················· | ····················· | |
| G | 3010 hCoV-19/United Arab Emirates/L2004034634/2020\|EPI_ | ···················· | ···················· | ····················· | ···················· | ···················· | ····················· | |

| | | | | | | | | |
|---|---|---|---|---|---|---|---|---|
| G | 3011 hCoV-19/United Arab Emirates/L2004034664/2020\|EPI_ | ···················· | ···················· | ···················· | ···················· | ···················· | ···················· | |
| G | 3012 hCoV-19/United Arab Emirates/L2004034680/2020\|EPI_ | ···················· | ···················· | ···················· | ···················· | ···················· | ···················· | |
| G | 3013 hCoV-19/United Arab Emirates/L2004035019/2020\|EPI_ | ···················· | ···················· | ···················· | ···················· | ···················· | ···················· | |
| G | 3014 hCoV-19/United Arab Emirates/L2004035133/2020\|EPI_ | ···················· | ···················· | ···················· | ···················· | ···················· | ···················· | |
| G | 3015 hCoV-19/United Arab Emirates/L2005036025/2020\|EPI_ | ···················· | ···················· | ···················· | ···················· | ···················· | ···················· | |
| G | 3016 hCoV-19/United Arab Emirates/L2005036083/2020\|EPI_ | ···················· | ···················· | ···················· | ···················· | ···················· | ···················· | |
| G | 3017 hCoV-19/Australia/VIC2195/2020\|EPI_ISL_521941\|2020 | ···················· | ···················· | ···················· | ···················· | ···················· | ···················· | |
| G | 3018 hCoV-19/Australia/VIC2196/2020\|EPI_ISL_521942\|2020 | ···················· | ···················· | ···················· | ···················· | ···················· | ···················· | |
| G | 3019 hCoV-19/Australia/VIC2197/2020\|EPI_ISL_521943\|2020 | ···················· | ···················· | ···················· | ···················· | ···················· | ···················· | |
| G | 3020 hCoV-19/Australia/VIC2198/2020\|EPI_ISL_521944\|2020 | ···················· | ···················· | ···················· | ···················· | ···················· | ···················· | |
| G | 3021 hCoV-19/Australia/VIC2202/2020\|EPI_ISL_521945\|2020 | ···················· | ···················· | ···················· | ···················· | ···················· | ···················· | |
| G | 3022 hCoV-19/Australia/VIC2204/2020\|EPI_ISL_521946\|2020 | ···················· | ···················· | ···················· | ···················· | ···················· | ···················· | |
| G | 3023 hCoV-19/Australia/VIC2205/2020\|EPI_ISL_521947\|2020 | ···················· | ···················· | ···················· | ···················· | ···················· | ···················· | |
| G | 3024 hCoV-19/Australia/VIC2206/2020\|EPI_ISL_521948\|2020 | ···················· | ···················· | ···················· | ···················· | ···················· | ···················· | |
| G | 3025 hCoV-19/Australia/VIC2210/2020\|EPI_ISL_521950\|2020 | ···················· | ···················· | ···················· | ···················· | ···················· | ···················· | |
| G | 3026 hCoV-19/Australia/VIC2211/2020\|EPI_ISL_521951\|2020 | ···················· | ···················· | ···················· | ···················· | ···················· | ···················· | |
| G | 3027 hCoV-19/Australia/VIC2212/2020\|EPI_ISL_521952\|2020 | ···················· | ···················· | ···················· | ···················· | ···················· | ···················· | |
| G | 3028 hCoV-19/Australia/VIC2213/2020\|EPI_ISL_521953\|2020 | ···················· | ···················· | ···················· | ···················· | ···················· | ···················· | |
| G | 3029 hCoV-19/Australia/VIC2214/2020\|EPI_ISL_521954\|2020 | ···················· | ···················· | ···················· | ···················· | ···················· | ···················· | |
| G | 3030 hCoV-19/Australia/VIC2216/2020\|EPI_ISL_521956\|2020 | ···················· | ···················· | ···················· | ···················· | ···················· | ···················· | |
| G | 3031 hCoV-19/Australia/VIC2217/2020\|EPI_ISL_521957\|2020 | ···················· | ···················· | ···················· | ···················· | ···················· | ···················· | |
| G | 3032 hCoV-19/Australia/VIC2220/2020\|EPI_ISL_521959\|2020 | ···················· | ···················· | ···················· | ···················· | ···················· | ···········M········ | |
| G | 3033 hCoV-19/Australia/VIC2224/2020\|EPI_ISL_521961\|2020 | ···················· | ···················· | ···················· | ···················· | ···················· | ···················· | |
| G | 3034 hCoV-19/Australia/VIC2226/2020\|EPI_ISL_521963\|2020 | ···················· | ···················· | ···················· | ···················· | ···················· | ···················· | |
| G | 3035 hCoV-19/Australia/VIC2227/2020\|EPI_ISL_521964\|2020 | ···················· | ···················· | ···················· | ···················· | ···················· | ···················· | |
| G | 3036 hCoV-19/Australia/VIC2228/2020\|EPI_ISL_521965\|2020 | ···················· | ·A·················· | ···················· | ···················· | ···················· | ···A················ | |
| G | 3037 hCoV-19/Australia/VIC2229/2020\|EPI_ISL_521966\|2020 | ···················· | ···················· | ···················· | ···················· | ···················· | ···················· | |
| G | 3038 hCoV-19/Australia/VIC2230/2020\|EPI_ISL_521967\|2020 | ···················· | ···················· | ···················· | ···················· | ···················· | ···················· | |
| G | 3039 hCoV-19/Australia/VIC2231/2020\|EPI_ISL_521968\|2020 | ···················· | ···················· | ···················· | ···················· | ···················· | ···················· | |
| G | 3040 hCoV-19/Australia/VIC2232/2020\|EPI_ISL_521969\|2020 | ···················· | ···················· | ···················· | ···················· | ···················· | ···················· | |
| G | 3041 hCoV-19/Australia/VIC2234/2020\|EPI_ISL_521971\|2020 | ···················· | ···················· | ···················· | ···················· | ···················· | ···················· | |
| G | 3048 hCoV-19/Australia/VIC2245/2020\|EPI_ISL_521978\|2020 | ···················· | ···················· | ···················· | ···················· | ···················· | ···················· | |
| G | 3050 hCoV-19/Australia/VIC2251/2020\|EPI_ISL_521982\|2020 | ···················· | ···················· | ···················· | ···················· | ···················· | ···················· | |
| G | 3051 hCoV-19/Australia/VIC2255/2020\|EPI_ISL_521984\|2020 | ···················· | ···················· | ···················· | ···················· | ···················· | ···················· | |
| G | 3052 hCoV-19/Australia/VIC2256/2020\|EPI_ISL_521985\|2020 | ···················· | ···················· | ···················· | ···················· | NNN················· | ···················· | 1 |
| G | 3053 hCoV-19/Australia/VIC2257/2020\|EPI_ISL_521986\|2020 | ···················· | ·A·················· | ···················· | ···················· | ···················· | ···A················ | |
| G | 3054 hCoV-19/Australia/VIC2260/2020\|EPI_ISL_521988\|2020 | ···················· | ···················· | ···················· | ···················· | NNN················· | ···················· | 1 |
| G | 3055 hCoV-19/Australia/VIC2261/2020\|EPI_ISL_521989\|2020 | ···················· | ···················· | ···················· | ···················· | ···················· | ···················· | |
| G | 3056 hCoV-19/Australia/VIC2269/2020\|EPI_ISL_521991\|2020 | ···················· | ···················· | ···················· | ···················· | ···················· | ···················· | |
| G | 3058 hCoV-19/Australia/VIC2271/2020\|EPI_ISL_521993\|2020 | ···················· | ···················· | ···················· | ···················· | ···················· | ···················· | |
| G | 3059 hCoV-19/Australia/VIC2272/2020\|EPI_ISL_521994\|2020 | ···················· | ···················· | ···················· | ···················· | ···················· | ···················· | |
| G | 3061 hCoV-19/Australia/VIC2274/2020\|EPI_ISL_521996\|2020 | ···················· | ···················· | ···················· | ···················· | ···················· | ···················· | |
| G | 3062 hCoV-19/Australia/VIC2275/2020\|EPI_ISL_521997\|2020 | ···················· | ···················· | ···················· | ···················· | ···················· | ···················· | |
| G | 3063 hCoV-19/Australia/VIC2276/2020\|EPI_ISL_521998\|2020 | ···················· | ···················· | ···················· | ···················· | ···················· | ···················· | |
| G | 3064 hCoV-19/Australia/VIC2277/2020\|EPI_ISL_521999\|2020 | ···················· | ···················· | ···················· | ···················· | ···················· | ···················· | |
| G | 3065 hCoV-19/Italy/ABR-IZSGC-TE46436/2020\|EPI_ISL_52900 | ···················· | ···················· | ···················· | ···················· | ···················· | ···················· | |
| G | 3066 hCoV-19/Italy/ABR-IZSGC-TE46439/2020\|EPI_ISL_52900 | ···················· | ···················· | ···················· | ···················· | ···················· | ···················· | |
| G | 3067 hCoV-19/Italy/ABR-IZSGC-TE49273/2020\|EPI_ISL_52900 | ···················· | ···················· | ···················· | ···················· | ···················· | ···················· | |
| G | 3068 hCoV-19/Italy/ABR-IZSGC-TE49480/2020\|EPI_ISL_52900 | ···················· | ···················· | ···················· | ···················· | ···················· | ···················· | |
| G | 3069 hCoV-19/Italy/ABR-IZSGC-TE49482/2020\|EPI_ISL_52900 | ···················· | ···················· | ···················· | ···················· | ···················· | ···················· | |
| G | 3070 hCoV-19/Italy/ABR-IZSGC-TE77497/2020\|EPI_ISL_52900 | ···················· | ···················· | ···················· | ···················· | ···················· | ···················· | |
| G | 3071 hCoV-19/Italy/ABR-IZSGC-TE79445/2020\|EPI_ISL_52900 | ···················· | ···················· | ···················· | ···················· | ···················· | ···················· | |
| G | 3072 hCoV-19/Italy/ABR-IZSGC-TE9047/2020\|EPI_ISL_529009 | ···················· | ···················· | ···················· | ···················· | ···················· | ···················· | |
| G | 3073 hCoV-19/Australia/VIC6110/2020\|EPI_ISL_518130\|2020 | ···················· | ···················· | ···················· | ···················· | ···················· | ···················· | |
| G | 3074 hCoV-19/Australia/VIC6111/2020\|EPI_ISL_518131\|2020 | ···················· | ···················· | ···················· | ···················· | ···················· | ···················· | |
| G | 3075 hCoV-19/Australia/VIC6113/2020\|EPI_ISL_518132\|2020 | ···················· | ···················· | ···················· | ···················· | ···················· | ···················· | |
| G | 3076 hCoV-19/Australia/VIC6114/2020\|EPI_ISL_518133\|2020 | ···················· | ···················· | ···················· | ···················· | ···················· | ···················· | |
| G | 3077 hCoV-19/Australia/VIC6116/2020\|EPI_ISL_518134\|2020 | ···················· | ···················· | ···················· | ···················· | ···················· | ···················· | |
| G | 3078 hCoV-19/Australia/VIC6117/2020\|EPI_ISL_518135\|2020 | ···················· | ···················· | ···················· | ···················· | ···················· | ···················· | |
| G | 3079 hCoV-19/Australia/VIC6118/2020\|EPI_ISL_518136\|2020 | ···················· | ···················· | ···················· | ···················· | ···················· | ···················· | |
| G | 3080 hCoV-19/Australia/VIC6120/2020\|EPI_ISL_518138\|2020 | ···················· | ···················· | ···················· | ···················· | ···················· | ···················· | |
| G | 3081 hCoV-19/Australia/VIC6121/2020\|EPI_ISL_518139\|2020 | ···················· | ···················· | ···················· | ···················· | ···················· | ···················· | |
| G | 3082 hCoV-19/Australia/VIC6123/2020\|EPI_ISL_518140\|2020 | ···················· | ···················· | ···················· | ···················· | ···················· | ···················· | |
| G | 3083 hCoV-19/Australia/VIC6126/2020\|EPI_ISL_518141\|2020 | ···················· | ···················· | ···················· | ···················· | ···················· | ···················· | |
| G | 3084 hCoV-19/Australia/VIC6128/2020\|EPI_ISL_518142\|2020 | ···················· | ···················· | ···················· | ···················· | ···················· | ···················· | |
| G | 3085 hCoV-19/Australia/VIC6129/2020\|EPI_ISL_518143\|2020 | ···················· | ···················· | ···················· | ···················· | ···················· | ···················· | |
| G | 3086 hCoV-19/Australia/VIC6131/2020\|EPI_ISL_518144\|2020 | ···················· | ···················· | ···················· | ···················· | ···················· | ···················· | |
| G | 3087 hCoV-19/Australia/VIC6132/2020\|EPI_ISL_518145\|2020 | ···················· | ···················· | ···················· | ···················· | ···················· | ···················· | |
| G | 3088 hCoV-19/Australia/VIC6133/2020\|EPI_ISL_518146\|2020 | ···················· | ···················· | ···················· | ···················· | ···················· | ···················· | |
| G | 3089 hCoV-19/Australia/VIC6135/2020\|EPI_ISL_518147\|2020 | ···················· | ···················· | ···················· | ···················· | ···················· | ···················· | |
| G | 3090 hCoV-19/Australia/VIC6137/2020\|EPI_ISL_518148\|2020 | ···················· | ···················· | ···················· | ···················· | ···················· | ···················· | |
| G | 3091 hCoV-19/Australia/VIC6138/2020\|EPI_ISL_518149\|2020 | ···················· | ···················· | ···················· | ···················· | ···················· | ···················· | |
| G | 3092 hCoV-19/Australia/VIC6139/2020\|EPI_ISL_518150\|2020 | ···················· | ···················· | ···················· | ···················· | ···················· | ···················· | |
| G | 3093 hCoV-19/Australia/VIC6140/2020\|EPI_ISL_518151\|2020 | ···················· | ···················· | ···················· | ···················· | ···················· | ···················· | |
| G | 3094 hCoV-19/Australia/VIC6141/2020\|EPI_ISL_518152\|2020 | ···················· | ···················· | ···················· | ···················· | ···················· | ···················· | |
| G | 3095 hCoV-19/Australia/VIC6142/2020\|EPI_ISL_518153\|2020 | ···················· | ···················· | ···················· | ···················· | ···················· | ···················· | |
| G | 3096 hCoV-19/Australia/VIC6143/2020\|EPI_ISL_518154\|2020 | ···················· | ···················· | ···················· | ···················· | ···················· | ···················· | |
| G | 3097 hCoV-19/Australia/VIC6144/2020\|EPI_ISL_518155\|2020 | ···················· | ···················· | ···················· | ···················· | ···················· | ···················· | |
| G | 3099 hCoV-19/Australia/VIC6146/2020\|EPI_ISL_518157\|2020 | ···················· | ···················· | ···················· | ···················· | ···················· | ···················· | |
| G | 3100 hCoV-19/Australia/VIC6147/2020\|EPI_ISL_518158\|2020 | ···················· | ···················· | ···················· | ···················· | ···················· | ···················· | |

| | | | | | | | | |
|---|---|---|---|---|---|---|---|---|
| G | 3101 | hCoV-19/Australia/VIC6148/2020\|EPI_ISL_518159\|2020 | .................... | .................... | ..................... | .................... | .................... | .................... |
| G | 3102 | hCoV-19/Australia/VIC6149/2020\|EPI_ISL_518160\|2020 | .................... | .................... | ..................... | .................... | .................... | .................... |
| G | 3103 | hCoV-19/Australia/VIC6150/2020\|EPI_ISL_518161\|2020 | .................... | .................... | ..................... | .................... | .................... | .................... |
| G | 3104 | hCoV-19/Australia/VIC6151/2020\|EPI_ISL_518162\|2020 | .................... | .................... | ..................... | .................... | .................... | .................... |
| G | 3105 | hCoV-19/Australia/VIC6154/2020\|EPI_ISL_518163\|2020 | .................... | .................... | ..................... | .................... | .................... | .................... |
| G | 3106 | hCoV-19/Australia/VIC6158/2020\|EPI_ISL_518165\|2020 | .................... | .................... | ..................... | .................... | .................... | .................... |
| G | 3107 | hCoV-19/Australia/VIC6159/2020\|EPI_ISL_518166\|2020 | .................... | .................... | ..................... | .................... | .................... | .................... |
| G | 3108 | hCoV-19/Australia/VIC6161/2020\|EPI_ISL_518167\|2020 | .................... | .................... | ..................... | .................... | .................... | .................... |
| G | 3109 | hCoV-19/Australia/VIC6163/2020\|EPI_ISL_518169\|2020 | .................... | .................... | ..................... | .................... | .................... | .................... |
| G | 3110 | hCoV-19/Australia/VIC6164/2020\|EPI_ISL_518170\|2020 | .................... | .................... | ..................... | .................... | .................... | .................... |
| G | 3111 | hCoV-19/Australia/VIC6165/2020\|EPI_ISL_518171\|2020 | .................... | .................... | ..................... | .................... | .................... | .................... |
| G | 3112 | hCoV-19/Australia/VIC6168/2020\|EPI_ISL_518174\|2020 | .................... | .................... | ..................... | .................... | .................... | .................... |
| G | 3113 | hCoV-19/Australia/VIC6169/2020\|EPI_ISL_518175\|2020 | .................... | .................... | ..................... | .................... | .................... | .................... |
| G | 3114 | hCoV-19/Australia/VIC6170/2020\|EPI_ISL_518176\|2020 | .................... | .................... | ..................... | .................... | .................... | .................... |
| G | 3115 | hCoV-19/Australia/VIC6171/2020\|EPI_ISL_518177\|2020 | .................... | .................... | ..................... | .................... | .................... | .................... |
| G | 3116 | hCoV-19/Australia/VIC6174/2020\|EPI_ISL_518179\|2020 | .................... | .................... | ..................... | .................... | .................... | .................... |
| G | 3117 | hCoV-19/Australia/VIC6175/2020\|EPI_ISL_518180\|2020 | .................... | .................... | ..................... | .................... | .................... | .................... |
| G | 3118 | hCoV-19/Australia/VIC6176/2020\|EPI_ISL_518181\|2020 | .................... | .................... | ..................... | .................... | .................... | .................... |
| G | 3119 | hCoV-19/Australia/VIC6177/2020\|EPI_ISL_518182\|2020 | .................... | .................... | ..................... | .................... | .................... | .................... |
| G | 3120 | hCoV-19/Australia/VIC6178/2020\|EPI_ISL_518183\|2020 | .................... | .................... | ..................... | .................... | .................... | .................... |
| G | 3121 | hCoV-19/Australia/VIC6179/2020\|EPI_ISL_518184\|2020 | .................... | .................... | ..................... | .................... | .................... | .................... |
| G | 3122 | hCoV-19/Australia/VIC6180/2020\|EPI_ISL_518185\|2020 | .................... | .................... | ..................... | .................... | .................... | .................... |
| G | 3123 | hCoV-19/Australia/VIC6181/2020\|EPI_ISL_518186\|2020 | .................... | .................... | ..................... | .................... | .................... | .................... |
| G | 3124 | hCoV-19/Australia/VIC6183/2020\|EPI_ISL_518188\|2020 | .................... | .................... | ..................... | .................... | .................... | .................... |
| G | 3125 | hCoV-19/Australia/VIC6188/2020\|EPI_ISL_518191\|2020 | .................... | .................... | ..................... | .................... | .................... | .................... |
| G | 3126 | hCoV-19/Australia/VIC6192/2020\|EPI_ISL_518194\|2020 | .................... | .................... | ..................... | .................... | .................... | .................... |
| G | 3127 | hCoV-19/Australia/VIC6193/2020\|EPI_ISL_518195\|2020 | .................... | .................... | ..................... | .................... | .................... | .................... |
| G | 3128 | hCoV-19/Australia/VIC6194/2020\|EPI_ISL_518196\|2020 | .................... | .................... | ..................... | .................... | .................... | .................... |
| G | 3129 | hCoV-19/Australia/VIC6196/2020\|EPI_ISL_518198\|2020 | .................... | .................... | ..................... | .................... | .................... | .................... |
| G | 3130 | hCoV-19/Australia/VIC6197/2020\|EPI_ISL_518199\|2020 | .................... | .................... | ..................... | .................... | .................... | .................... |
| G | 3131 | hCoV-19/Australia/VIC4084/2020\|EPI_ISL_519465\|2020 | .................... | .................... | ..................... | .................... | .................... | .................... |
| G | 3132 | hCoV-19/Australia/VIC4089/2020\|EPI_ISL_519470\|2020 | .................... | .................... | ..................... | .................... | .................... | .................... |
| G | 3133 | hCoV-19/Australia/VIC4090/2020\|EPI_ISL_519471\|2020 | .................... | .................... | ..................... | .................... | .................... | .................... |
| G | 3134 | hCoV-19/Australia/VIC4091/2020\|EPI_ISL_519472\|2020 | .................... | .................... | ..................... | .................... | .................... | .................... |
| G | 3135 | hCoV-19/Australia/VIC4092/2020\|EPI_ISL_519473\|2020 | .................... | .................... | ..................... | .................... | .................... | .................... |
| G | 3136 | hCoV-19/Australia/VIC4093/2020\|EPI_ISL_519474\|2020 | .................... | .................... | ..................... | .................... | .................... | .................... |
| G | 3137 | hCoV-19/Australia/VIC4095/2020\|EPI_ISL_519476\|2020 | .................... | .................... | ..................... | .................... | .................... | .................... |
| G | 3138 | hCoV-19/Australia/VIC4097/2020\|EPI_ISL_519478\|2020 | .................... | .................... | ..................... | .................... | .................... | .................... |
| G | 3139 | hCoV-19/Australia/VIC4100/2020\|EPI_ISL_519481\|2020 | .................... | .................... | ..................... | .................... | .................... | .................... |
| G | 3140 | hCoV-19/Australia/VIC4101/2020\|EPI_ISL_519482\|2020 | .................... | .................... | ..................... | .................... | .................... | .................... |
| G | 3141 | hCoV-19/Australia/VIC4104/2020\|EPI_ISL_519484\|2020 | .................... | .................... | ..................... | .................... | .................... | .................... |
| G | 3142 | hCoV-19/Australia/VIC4105/2020\|EPI_ISL_519485\|2020 | .................... | .................... | ..................... | .................... | .................... | .................... |
| G | 3143 | hCoV-19/Australia/VIC4323/2020\|EPI_ISL_519620\|2020 | .................... | .................... | ..................... | .................... | .................... | .................... |
| G | 3144 | hCoV-19/Australia/VIC4324/2020\|EPI_ISL_519621\|2020 | .................... | .................... | ..................... | .................... | .................... | .................... |
| G | 3145 | hCoV-19/Australia/VIC4325/2020\|EPI_ISL_519622\|2020 | .................... | .................... | ..................... | .................... | .................... | .................... |
| G | 3146 | hCoV-19/Australia/VIC4326/2020\|EPI_ISL_519623\|2020 | .................... | .................... | ..................... | .................... | .................... | .................... |
| G | 3147 | hCoV-19/Australia/VIC4328/2020\|EPI_ISL_519625\|2020 | .................... | .................... | ..................... | .................... | .................... | .................... |
| G | 3148 | hCoV-19/Australia/VIC4330/2020\|EPI_ISL_519627\|2020 | .................... | .................... | ..................... | .................... | .................... | .................... |
| G | 3149 | hCoV-19/Australia/VIC4331/2020\|EPI_ISL_519628\|2020 | .................... | .................... | ..................... | .................... | .................... | .................... |
| G | 3150 | hCoV-19/Australia/VIC4333/2020\|EPI_ISL_519629\|2020 | .................... | .................... | ..................... | .................... | .................... | .................... |
| G | 3151 | hCoV-19/Australia/VIC4337/2020\|EPI_ISL_519633\|2020 | .................... | .................... | ..................... | .................... | .................... | .................... |
| G | 3152 | hCoV-19/Australia/VIC3716/2020\|EPI_ISL_520449\|2020 | .................... | .................... | ..................... | .................... | .................... | .................... |
| G | 3153 | hCoV-19/Australia/VIC3887/2020\|EPI_ISL_520600\|2020 | .................... | .................... | ..................... | .................... | .................... | .................... |
| G | 3154 | hCoV-19/Australia/VIC3889/2020\|EPI_ISL_520601\|2020 | .................... | .................... | ..................... | .................... | .................... | .................... |
| G | 3155 | hCoV-19/Australia/VIC3890/2020\|EPI_ISL_520602\|2020 | .................... | .................... | ..................... | .................... | .................... | .................... |
| G | 3156 | hCoV-19/Australia/VIC3891/2020\|EPI_ISL_520603\|2020 | .................... | .................... | ..................... | .................... | .................... | .................... |
| G | 3157 | hCoV-19/Australia/VIC3894/2020\|EPI_ISL_520606\|2020 | .................... | .................... | ..................... | .................... | .................... | .................... |
| G | 3159 | hCoV-19/Australia/VIC3896/2020\|EPI_ISL_520608\|2020 | .................... | .................... | ..................... | .................... | .................... | .................... |
| G | 3160 | hCoV-19/Australia/VIC3897/2020\|EPI_ISL_520609\|2020 | .................... | .................... | ..................... | .................... | .................... | .................... |
| G | 3161 | hCoV-19/Australia/VIC2946/2020\|EPI_ISL_521776\|2020 | .................... | .................... | ..................... | .................... | .................... | .................... |
| G | 3162 | hCoV-19/Australia/VIC2947/2020\|EPI_ISL_521777\|2020 | .................... | .................... | ..................... | .................... | .................... | .................... |
| G | 3164 | hCoV-19/Australia/VIC2949/2020\|EPI_ISL_521779\|2020 | .................... | .................... | ..................... | .................... | .................... | .................... |
| G | 3165 | hCoV-19/Australia/VIC2950/2020\|EPI_ISL_521780\|2020 | .................... | .................... | ..................... | .................... | .................... | .................... |
| G | 3166 | hCoV-19/Australia/VIC2951/2020\|EPI_ISL_521781\|2020 | .................... | .................... | ..................... | .................... | .................... | .................... |
| G | 3167 | hCoV-19/Australia/VIC2952/2020\|EPI_ISL_521782\|2020 | .................... | .................... | ..................... | .................... | .................... | .................... |
| G | 3168 | hCoV-19/Australia/VIC2953/2020\|EPI_ISL_521783\|2020 | .................... | .................... | ..................... | .................... | .................... | .................... |
| G | 3169 | hCoV-19/Australia/VIC2954/2020\|EPI_ISL_521784\|2020 | .................... | .................... | ..................... | .................... | .................... | .................... |
| G | 3170 | hCoV-19/Australia/VIC2955/2020\|EPI_ISL_521785\|2020 | .................... | .................... | ..................... | .................... | .................... | .................... |
| G | 3171 | hCoV-19/Australia/VIC2956/2020\|EPI_ISL_521786\|2020 | .................... | .................... | ..................... | .................... | .................... | .................... |
| G | 3172 | hCoV-19/Australia/VIC2957/2020\|EPI_ISL_521787\|2020 | .................... | .................... | ..................... | .................... | .................... | .................... |
| G | 3174 | hCoV-19/Australia/VIC2959/2020\|EPI_ISL_521789\|2020 | .................... | .................... | ..................... | .................... | .................... | .................... |
| G | 3175 | hCoV-19/Australia/VIC2960/2020\|EPI_ISL_521790\|2020 | .................... | .................... | ..................... | .................... | .................... | .................... |
| G | 3176 | hCoV-19/Australia/VIC2961/2020\|EPI_ISL_521791\|2020 | .................... | .................... | ..................... | .................... | .................... | .................... |
| G | 3177 | hCoV-19/Australia/VIC2962/2020\|EPI_ISL_521792\|2020 | .................... | .................... | ..................... | .................... | .................... | .................... |
| G | 3178 | hCoV-19/Australia/VIC2963/2020\|EPI_ISL_521793\|2020 | .................... | .................... | ..................... | .................... | .................... | .................... |
| G | 3179 | hCoV-19/Australia/VIC2965/2020\|EPI_ISL_521795\|2020 | .................... | .................... | ..................... | .................... | .................... | .................... |
| G | 3180 | hCoV-19/Australia/VIC2966/2020\|EPI_ISL_521796\|2020 | .................... | .................... | ..................... | .................... | .................... | .................... |
| G | 3181 | hCoV-19/Australia/VIC2967/2020\|EPI_ISL_521797\|2020 | .................... | .................... | ..................... | .................... | .................... | .................... |
| G | 3182 | hCoV-19/Australia/VIC2968/2020\|EPI_ISL_521798\|2020 | .................... | .................... | ..................... | .................... | .................... | .................... |
| G | 3183 | hCoV-19/Australia/VIC2969/2020\|EPI_ISL_521799\|2020 | .................... | .................... | ..................... | .................... | .................... | .................... |

```
G    3185 hCoV-19/Australia/VIC2185/2020|EPI_ISL_521931|2020    ····················  ····················  ·····················  ····················  ····················  ····················
G    3187 hCoV-19/Australia/VIC2187/2020|EPI_ISL_521933|2020    ····················  ····················  ·····················  ····················  ····················  ····················
G    3188 hCoV-19/Australia/VIC2188/2020|EPI_ISL_521934|2020    ····················  ····················  ·····················  ····················  ····················  ····················
G    3189 hCoV-19/Australia/VIC2189/2020|EPI_ISL_521935|2020    ····················  ····················  ·····················  ····················  ····················  ····················
G    3190 hCoV-19/Australia/VIC2190/2020|EPI_ISL_521936|2020    ····················  ····················  ·····················  ····················  ····················  ····················
G    3191 hCoV-19/Australia/VIC2191/2020|EPI_ISL_521937|2020    ····················  ····················  ·····················  ····················  ····················  ····················
G    3192 hCoV-19/Australia/VIC2192/2020|EPI_ISL_521938|2020    ····················  ····················  ·····················  ····················  ····················  ····················
G    3193 hCoV-19/Australia/VIC2193/2020|EPI_ISL_521939|2020    ····················  ····················  ·····················  ····················  ····················  ····················
G    3194 hCoV-19/Australia/VIC2194/2020|EPI_ISL_521940|2020    ····················  ····················  ·····················  ····················  ····················  ····················
G    3195 hCoV-19/Russia/SPE-RII-12083V/2020|EPI_ISL_524000|    ····················  ····················  ·····················  ····················  ····················  ····················
G    3196 hCoV-19/Russia/SPE-RII-12101V/2020|EPI_ISL_524001|    ····················  ····················  ·····················  ····················  ····················  ····················
G    3197 hCoV-19/Russia/SPE-RII-12105V/2020|EPI_ISL_524002|    ····················  ····················  ·····················  ····················  ····················  ····················
G    3198 hCoV-19/Russia/SPE-RII-13179V/2020|EPI_ISL_524003|    ····················  ····················  ·····················  ····················  ····················  ····················
G    3199 hCoV-19/Russia/SPE-RII-13247V/2020|EPI_ISL_524004|    ····················  ····················  ·····················  ····················  ····················  ····················
G    3200 hCoV-19/Russia/SPE-RII-13482V/2020|EPI_ISL_524005|    ····················  ····················  ·····················  ····················  ····················  ····················
G    3201 hCoV-19/Russia/SPE-RII-13784V/2020|EPI_ISL_524006|    ····················  ····················  ·····················  ····················  ····················  ····················
G    3202 hCoV-19/Russia/SPE-RII-14716V/2020|EPI_ISL_524007|    ····················  ····················  ·····················  ····················  ····················  ····················
G    3203 hCoV-19/Russia/SPE-RII-15054S/2020|EPI_ISL_524008|    ····················  ····················  ·····················  ····················  ····················  ····················
G    3204 hCoV-19/USA/UT-08287/2020|EPI_ISL_524086|2020-05-2    ····················  ····················  ·····················  ····················  ····················  ····················
G    3205 hCoV-19/USA/UT-UPHL-2008983/2020|EPI_ISL_525300|20    ····················  ····················  ·····················  ····················  ····················  ····················
G    3206 hCoV-19/USA/UT-UPHL-2008985/2020|EPI_ISL_525302|20    ····················  ····················  ·····················  ····················  ····················  ····················
G    3207 hCoV-19/USA/UT-UPHL-2008990/2020|EPI_ISL_525304|20    ····················  ····················  ·····················  ····················  ····················  ····················
G    3208 hCoV-19/USA/UT-UPHL-2008991/2020|EPI_ISL_525305|20    ····················  ····················  ·····················  ····················  ····················  ····················
G    3209 hCoV-19/USA/UT-UPHL-2008993/2020|EPI_ISL_525306|20    ····················  ····················  ·····················  ····················  ····················  ····················
G    3210 hCoV-19/USA/UT-UPHL-2008994/2020|EPI_ISL_525307|20    ····················  ····················  ·····················  ····················  ····················  ····················
G    3211 hCoV-19/USA/UT-UPHL-2008998/2020|EPI_ISL_525309|20    ····················  ····················  ·····················  ····················  ····················  ····················
G    3212 hCoV-19/USA/UT-UPHL-2008999/2020|EPI_ISL_525310|20    ····················  ····················  ·····················  ····················  ····················  ····················
G    3213 hCoV-19/USA/UT-UPHL-2009000/2020|EPI_ISL_525311|20    ····················  ····················  ·····················  ····················  ····················  ····················
G    3214 hCoV-19/USA/UT-UPHL-2009001/2020|EPI_ISL_525312|20    ····················  ····················  ·····················  ····················  ····················  ····················
G    3215 hCoV-19/USA/UT-UPHL-2009003/2020|EPI_ISL_525313|20    ····················  ····················  ·····················  ····················  ····················  ····················
G    3216 hCoV-19/USA/UT-UPHL-2009006/2020|EPI_ISL_525316|20    ····················  ····················  ·····················  ····················  ·········A··········  ····················
G    3217 hCoV-19/USA/UT-UPHL-2009008/2020|EPI_ISL_525317|20    ····················  ····················  ·····················  ····················  ····················  ····················
G    3218 hCoV-19/USA/UT-UPHL-2009011/2020|EPI_ISL_525319|20    ····················  ····················  ·····················  ····················  ····················  ····················
G    3219 hCoV-19/USA/UT-UPHL-2009013/2020|EPI_ISL_525320|20    ····················  ····················  ·····················  ····················  ····················  ····················
G    3220 hCoV-19/USA/UT-UPHL-2009015/2020|EPI_ISL_525321|20    ····················  ····················  ·····················  ····················  ····················  ····················
G    3221 hCoV-19/USA/UT-UPHL-2009016/2020|EPI_ISL_525322|20    ····················  ····················  ·····················  ····················  ····················  ····················
G    3222 hCoV-19/USA/UT-UPHL-2009019/2020|EPI_ISL_525325|20    ····················  ····················  ·····················  ····················  ····················  ····················
G    3223 hCoV-19/USA/UT-UPHL-2009020/2020|EPI_ISL_525326|20    ····················  ····················  ·····················  ····················  ····················  ····················
G    3224 hCoV-19/USA/UT-UPHL-2009022/2020|EPI_ISL_525328|20    ····················  ····················  ·····················  ····················  ····················  ····················
G    3225 hCoV-19/USA/UT-UPHL-2009024/2020|EPI_ISL_525330|20    ····················  ····················  ·····················  ····················  ····················  ····················
G    3226 hCoV-19/USA/UT-UPHL-2009029/2020|EPI_ISL_525334|20    ····················  ····················  ·····················  ····················  ····················  ····················
G    3227 hCoV-19/USA/UT-UPHL-04471/2020|EPI_ISL_525338|2020    ····················  ····················  ·····················  ····················  ····················  ····················
G    3228 hCoV-19/USA/UT-UPHL-04472/2020|EPI_ISL_525339|2020    ····················  ····················  ·····················  ····················  ····················  ····················
G    3229 hCoV-19/USA/FL-BPHL-1114/2020|EPI_ISL_526600|2020-    ····················  ····················  ·····················  ····················  ····················  ····················
G    3230 hCoV-19/USA/FL-BPHL-1116/2020|EPI_ISL_526602|2020-    ····················  ·A··················  ·····················  ····················  ····················  ····················
G    3231 hCoV-19/USA/FL-BPHL-1117/2020|EPI_ISL_526603|2020-    ····················  ·A··················  ·····················  ····················  ····················  ····················
G    3232 hCoV-19/USA/FL-BPHL-1119/2020|EPI_ISL_526605|2020-    ····················  ····················  ·····················  ····················  ····················  ····················
G    3233 hCoV-19/USA/FL-BPHL-1120/2020|EPI_ISL_526606|2020-    ····················  ····················  ·····················  ····················  ····················  ····················
G    3234 hCoV-19/USA/FL-BPHL-1121/2020|EPI_ISL_526607|2020-    ····················  ····················  ·····················  ····················  ····················  ····················
G    3235 hCoV-19/USA/FL-BPHL-1122/2020|EPI_ISL_526608|2020-    ····················  ····················  ·····················  ····················  ····················  ····················
G    3236 hCoV-19/USA/FL-BPHL-1124/2020|EPI_ISL_526610|2020-    ····················  ····················  ·····················  ····················  ····················  ····················
G    3237 hCoV-19/USA/FL-BPHL-1125/2020|EPI_ISL_526611|2020-    ····················  ····················  ·····················  ····················  ····················  ····················
G    3239 hCoV-19/USA/FL-BPHL-1127/2020|EPI_ISL_526613|2020-    ····················  ····················  ·····················  ····················  ····················  ····················
G    3240 hCoV-19/USA/FL-BPHL-1128/2020|EPI_ISL_526614|2020-    ····················  ····················  ·····················  ····················  ····················  ····················
G    3241 hCoV-19/USA/FL-BPHL-1130/2020|EPI_ISL_526616|2020-    ····················  ····················  ·····················  ····················  ····················  ····················
G    3242 hCoV-19/USA/FL-BPHL-1131/2020|EPI_ISL_526617|2020-    ····················  ····················  ·····················  ····················  ····················  ····················
G    3243 hCoV-19/USA/FL-BPHL-1132/2020|EPI_ISL_526618|2020-    ····················  ····················  ·····················  ····················  ····················  ····················
G    3244 hCoV-19/USA/FL-BPHL-1133/2020|EPI_ISL_526619|2020-    ····················  ····················  ·····················  ····················  ····················  ····················
G    3245 hCoV-19/USA/FL-BPHL-1134/2020|EPI_ISL_526620|2020-    ····················  ····················  ·····················  ····················  ····················  ····················
G    3246 hCoV-19/USA/FL-BPHL-1135/2020|EPI_ISL_526621|2020-    ····················  ····················  ·····················  ····················  ····················  ····················
G    3247 hCoV-19/USA/FL-BPHL-1136/2020|EPI_ISL_526622|2020-    ····················  ····················  ·····················  ····················  ····················  ····················
G    3248 hCoV-19/USA/FL-BPHL-1137/2020|EPI_ISL_526623|2020-    ····················  ····················  ·····················  ····················  ····················  ····················
G    3249 hCoV-19/USA/FL-BPHL-1138/2020|EPI_ISL_526624|2020-    ····················  ····················  ·····················  ····················  ····················  ····················
G    3250 hCoV-19/USA/FL-BPHL-1139/2020|EPI_ISL_526625|2020-    ····················  ····················  ·····················  ····················  ····················  ····················
G    3251 hCoV-19/USA/FL-BPHL-1140/2020|EPI_ISL_526626|2020-    ····················  ····················  ·····················  ····················  ····················  ····················
G    3252 hCoV-19/USA/FL-BPHL-1141/2020|EPI_ISL_526627|2020-    ····················  ····················  ·····················  ····················  ····················  ····················
G    3253 hCoV-19/USA/FL-BPHL-1142/2020|EPI_ISL_526628|2020-    ····················  ····················  ·····················  ····················  ····················  ····················
G    3254 hCoV-19/USA/FL-BPHL-1143/2020|EPI_ISL_526629|2020-    ····················  ····················  ·····················  ····················  ····················  ····················
G    3255 hCoV-19/USA/FL-BPHL-1144/2020|EPI_ISL_526630|2020-    ····················  ····················  ·····················  ····················  ····················  ····················
G    3256 hCoV-19/USA/FL-BPHL-1145/2020|EPI_ISL_526631|2020-    ····················  ····················  ·····················  ····················  ····················  ····················
G    3257 hCoV-19/USA/FL-BPHL-1146/2020|EPI_ISL_526632|2020-    ····················  ····················  ·····················  ····················  ····················  ····················
G    3258 hCoV-19/USA/FL-BPHL-1147/2020|EPI_ISL_526633|2020-    ····················  ····················  ·····················  ····················  ····················  ····················
G    3260 hCoV-19/USA/FL-BPHL-1149/2020|EPI_ISL_526635|2020-    ····················  ····················  ·····················  ····················  ····················  ····················
G    3262 hCoV-19/USA/FL-BPHL-1151/2020|EPI_ISL_526637|2020-    ····················  ····················  ·····················  ····················  ····················  ····················
G    3263 hCoV-19/USA/FL-BPHL-1152/2020|EPI_ISL_526638|2020-    ····················  ····················  ·····················  ····················  ····················  ····················
G    3264 hCoV-19/USA/FL-BPHL-1157/2020|EPI_ISL_526643|2020-    ····················  ····················  ·····················  ····················  ····················  ····················
G    3265 hCoV-19/USA/FL-BPHL-1158/2020|EPI_ISL_526644|2020-    ····················  ····················  ·····················  ····················  ····················  ····················
G    3266 hCoV-19/USA/FL-BPHL-1159/2020|EPI_ISL_526645|2020-    ····················  ····················  ·····················  ····················  ····················  ····················
G    3267 hCoV-19/USA/FL-BPHL-1160/2020|EPI_ISL_526646|2020-    ····················  ····················  ·····················  ····················  ····················  ····················
G    3269 hCoV-19/USA/FL-BPHL-1163/2020|EPI_ISL_526649|2020-    ····················  ····················  ·····················  ····················  ····················  ····················
```

G 3271 hCoV-19/USA/FL-BPHL-1166/2020|EPI_ISL_526652|2020- ···················· ···················· ····················· ···················· ···················· ····················
G 3272 hCoV-19/USA/FL-BPHL-1167/2020|EPI_ISL_526653|2020- ···················· ···················· ····················· ···················· ···················· ····················
G 3275 hCoV-19/USA/FL-BPHL-1174/2020|EPI_ISL_526660|2020- ···················· ···················· ····················· ···················· ···················· ····················
G 3276 hCoV-19/USA/FL-BPHL-1175/2020|EPI_ISL_526661|2020- ···················· ···················· ····················· ···················· ···················· ····················
G 3277 hCoV-19/USA/FL-BPHL-1176/2020|EPI_ISL_526662|2020- ···················· ···················· ····················· ···················· ···················· ····················
G 3278 hCoV-19/USA/FL-BPHL-1177/2020|EPI_ISL_526663|2020- ···················· ···················· ····················· ···················· ···················· ····················
G 3279 hCoV-19/USA/FL-BPHL-1178/2020|EPI_ISL_526664|2020- ···················· ···················· ····················· ···················· ···················· ····················
G 3280 hCoV-19/USA/FL-BPHL-1179/2020|EPI_ISL_526665|2020- ···················· ···················· ····················· ···················· ···················· ····················
G 3281 hCoV-19/USA/FL-BPHL-1180/2020|EPI_ISL_526666|2020- ···················· ···················· ····················· ···················· ···················· ····················
G 3282 hCoV-19/USA/FL-BPHL-1181/2020|EPI_ISL_526667|2020- ···················· ···················· ····················· ···················· ···················· ····················
G 3283 hCoV-19/USA/FL-BPHL-1183/2020|EPI_ISL_526669|2020- ···················· ···················· ····················· ···················· ···················· ····················
G 3284 hCoV-19/Nigeria/OY605-CV170/2020|EPI_ISL_527901|20 ···················· ···················· ····················· ···················· ······T············· ····················
G 3285 hCoV-19/Nigeria/OY608-CV172/2020|EPI_ISL_527903|20 ···················· ···················· ····················· ···················· ···················· ····················
G 3286 hCoV-19/Nigeria/OY612-CV173/2020|EPI_ISL_527904|20 ···················· ···················· ····················· ···················· ···················· ····················
G 3287 hCoV-19/Nigeria/OY616-CV174/2020|EPI_ISL_527905|20 ···················· ···················· ····················· ···················· ······T············· ····················
G 3288 hCoV-19/Nigeria/OY626-CV183/2020|EPI_ISL_527910|20 ···················· ···················· ····················· ···················· ···················· ····················
G 3289 hCoV-19/Nigeria/OY627-CV184/2020|EPI_ISL_527911|20 ···················· ···················· ····················· ···················· ···················· ····················
G 3290 hCoV-19/Nigeria/OY628-CV185/2020|EPI_ISL_527912|20 ········T··········· ···················· ····················· ···················· ···················· ····················
G 3291 hCoV-19/Nigeria/OY633-CV189/2020|EPI_ISL_527914|20 ···················· ···················· ····················· ···················· ···················· ····················
G 3292 hCoV-19/Nigeria/OY636-CV192/2020|EPI_ISL_527915|20 ···················· ···················· ····················· ···················· ···················· ····················
G 3293 hCoV-19/Switzerland/BS-UHB-42167340/2020|EPI_ISL_5 ···················· ···················· ····················· ···················· ···················· ····················
G 3294 hCoV-19/Switzerland/BL-UHB-42167842/2020|EPI_ISL_5 ···················· ···················· ····················· ···················· ···················· ····················
G 3296 hCoV-19/Switzerland/BS-UHB-42176753/2020|EPI_ISL_5 ···················· ···················· ····················· ···················· ···················· ····················
G 3297 hCoV-19/Switzerland/BL-UHB-42176771/2020|EPI_ISL_5 ···················· ···················· ····················· ···················· ···················· ····················
G 3299 hCoV-19/Switzerland/BS-UHB-42177239/2020|EPI_ISL_5 ···················· ···················· ····················· ···················· ·NNN················ ···················· 1
G 3300 hCoV-19/Switzerland/BS-UHB-42177430/2020|EPI_ISL_5 ···················· ···················· ····················· ···················· ···················· ····················
G 3301 hCoV-19/Switzerland/BS-UHB-42177434/2020|EPI_ISL_5 ···················· ···················· ····················· ···················· ···················· ····················
G 3302 hCoV-19/Switzerland/BS-UHB-42177585/2020|EPI_ISL_5 ···················· ···················· ····················· ···················· ···················· ····················
G 3306 hCoV-19/Switzerland/BS-UHB-42181327/2020|EPI_ISL_5 ···················· ···················· ····················· ···················· ···················· ····················
G 3307 hCoV-19/Switzerland/BS-UHB-42181461/2020|EPI_ISL_5 ···················· ···················· ····················· ···················· ···················· ····················
G 3318 hCoV-19/Switzerland/BS-UHB-42184365/2020|EPI_ISL_5 ···················· ···················· ····················· ···················· ·NN················· ····················
G 3320 hCoV-19/Switzerland/BL-UHB-42185206/2020|EPI_ISL_5 ···················· ···················· ····················· ···················· ···················· ····················
G 3329 hCoV-19/Australia/VIC6212/2020|EPI_ISL_518210|2020 ···················· ···················· ····················· ···················· ···················· ····················
G 3330 hCoV-19/Australia/VIC6213/2020|EPI_ISL_518211|2020 ···················· ···················· ····················· ···················· ···················· ····················
G 3331 hCoV-19/Australia/VIC6214/2020|EPI_ISL_518212|2020 ···················· ···················· ····················· ···················· ···················· ····················
G 3332 hCoV-19/Australia/VIC6215/2020|EPI_ISL_518213|2020 ···················· ···················· ····················· ···················· ···················· ····················
G 3333 hCoV-19/Australia/VIC6216/2020|EPI_ISL_518214|2020 ···················· ···················· ····················· ···················· ···················· ····················
G 3334 hCoV-19/Australia/VIC6222/2020|EPI_ISL_518220|2020 ···················· ···················· ····················· ···················· ···················· ····················
G 3335 hCoV-19/Australia/VIC6223/2020|EPI_ISL_518221|2020 ···················· ···················· ····················· ···················· ···················· ····················
G 3336 hCoV-19/Australia/VIC6225/2020|EPI_ISL_518222|2020 ···················· ···················· ····················· ···················· ···················· ····················
G 3337 hCoV-19/Australia/VIC6226/2020|EPI_ISL_518223|2020 ···················· ···················· ····················· ···················· ···················· ····················
G 3338 hCoV-19/Australia/VIC6227/2020|EPI_ISL_518224|2020 ···················· ···················· ····················· ···················· ···················· ····················
G 3339 hCoV-19/Australia/VIC6230/2020|EPI_ISL_518226|2020 ···················· ···················· ····················· ···················· ···················· ····················
G 3340 hCoV-19/Australia/VIC6231/2020|EPI_ISL_518227|2020 ···················· ···················· ····················· ···················· ···················· ····················
G 3341 hCoV-19/Australia/VIC6232/2020|EPI_ISL_518228|2020 ···················· ···················· ····················· ···················· ···················· ····················
G 3342 hCoV-19/Australia/VIC6235/2020|EPI_ISL_518229|2020 ···················· ···················· ····················· ···················· ···················· ····················
G 3343 hCoV-19/Australia/VIC6236/2020|EPI_ISL_518230|2020 ···················· ···················· ····················· ···················· ···················· ····················
G 3344 hCoV-19/Australia/VIC6237/2020|EPI_ISL_518231|2020 ···················· ···················· ····················· ···················· ···················· ····················
G 3345 hCoV-19/Australia/VIC6238/2020|EPI_ISL_518232|2020 ···················· ···················· ····················· ···················· ···················· ····················
G 3346 hCoV-19/Australia/VIC6240/2020|EPI_ISL_518234|2020 ···················· ···················· ····················· ···················· ···················· ····················
G 3347 hCoV-19/Australia/VIC6241/2020|EPI_ISL_518235|2020 ···················· ···················· ····················· ···················· ···················· ····················
G 3348 hCoV-19/Australia/VIC6242/2020|EPI_ISL_518236|2020 ···················· ···················· ····················· ···················· ···················· ····················
G 3349 hCoV-19/Australia/VIC6244/2020|EPI_ISL_518238|2020 ···················· ···················· ····················· ···················· ···················· ····················
G 3350 hCoV-19/Australia/VIC6246/2020|EPI_ISL_518239|2020 ···················· ···················· ····················· ···················· ···················· ····················
G 3351 hCoV-19/Australia/VIC6247/2020|EPI_ISL_518240|2020 ···················· ···················· ····················· ···················· ···················· ····················
G 3352 hCoV-19/Australia/VIC6248/2020|EPI_ISL_518241|2020 ···················· ···················· ····················· ···················· ···················· ····················
G 3353 hCoV-19/Australia/VIC6249/2020|EPI_ISL_518242|2020 ···················· ···················· ····················· ···················· ···················· ····················
G 3354 hCoV-19/Australia/VIC6251/2020|EPI_ISL_518244|2020 ···················· ···················· ····················· ···················· ···················· ····················
G 3355 hCoV-19/Australia/VIC6254/2020|EPI_ISL_518246|2020 ···················· ···················· ····················· ···················· ···················· ····················
G 3356 hCoV-19/Australia/VIC6255/2020|EPI_ISL_518247|2020 ···················· ···················· ····················· ···················· ···················· ····················
G 3357 hCoV-19/Australia/VIC6256/2020|EPI_ISL_518248|2020 ···················· ···················· ····················· ···················· ···················· ····················
G 3358 hCoV-19/Australia/VIC6257/2020|EPI_ISL_518249|2020 ···················· ···················· ····················· ···················· ···················· ····················
G 3359 hCoV-19/Australia/VIC4218/2020|EPI_ISL_519546|2020 ···················· ···················· ····················· ···················· ···················· ····················
G 3360 hCoV-19/Australia/VIC4219/2020|EPI_ISL_519547|2020 ···················· ···················· ····················· ···················· ···················· ····················
G 3361 hCoV-19/Australia/VIC4220/2020|EPI_ISL_519548|2020 ···················· ···················· ····················· ···················· ···················· ····················
G 3362 hCoV-19/Australia/VIC4221/2020|EPI_ISL_519549|2020 ···················· ···················· ····················· ···················· ···················· ····················
G 3363 hCoV-19/Australia/VIC4223/2020|EPI_ISL_519551|2020 ···················· ···················· ····················· ···················· ···················· ····················
G 3364 hCoV-19/Australia/VIC4224/2020|EPI_ISL_519552|2020 ···················· ···················· ····················· ···················· ···················· ····················
G 3365 hCoV-19/Australia/VIC4225/2020|EPI_ISL_519553|2020 ···················· ···················· ····················· ···················· ···················· ····················
G 3366 hCoV-19/Australia/VIC4226/2020|EPI_ISL_519554|2020 ···················· ···················· ····················· ···················· ···················· ····················
G 3367 hCoV-19/Australia/VIC4227/2020|EPI_ISL_519555|2020 ···················· ···················· ····················· ···················· ···················· ····················
G 3368 hCoV-19/Australia/VIC4228/2020|EPI_ISL_519556|2020 ···················· ···················· ····················· ···················· ···················· ····················
G 3369 hCoV-19/Australia/VIC4229/2020|EPI_ISL_519557|2020 ···················· ···················· ····················· ···················· ···················· ····················
G 3370 hCoV-19/Australia/VIC4230/2020|EPI_ISL_519558|2020 ···················· ···················· ····················· ···················· ···················· ····················
G 3371 hCoV-19/Australia/VIC4231/2020|EPI_ISL_519559|2020 ···················· ···················· ····················· ···················· ···················· ····················
G 3372 hCoV-19/Australia/VIC4232/2020|EPI_ISL_519560|2020 ···················· ···················· ····················· ···················· ···················· ····················
G 3373 hCoV-19/Australia/VIC4233/2020|EPI_ISL_519561|2020 ···················· ···················· ····················· ···················· ···················· ····················
G 3374 hCoV-19/Australia/VIC4234/2020|EPI_ISL_519562|2020 ···················· ···················· ····················· ···················· ···················· ····················
G 3375 hCoV-19/Australia/VIC4235/2020|EPI_ISL_519563|2020 ···················· ···················· ····················· ···················· ···················· ····················
G 3376 hCoV-19/Australia/VIC4236/2020|EPI_ISL_519564|2020 ···················· ···················· ····················· ···················· ···················· ····················

| | | | | | | | |
|---|---|---|---|---|---|---|---|
| G | 3377 hCoV-19/Australia/VIC4237/2020\|EPI_ISL_519565\|2020 | ···················· | ···················· | ···················· | ···················· | ···················· | ···················· |
| G | 3378 hCoV-19/Australia/VIC4238/2020\|EPI_ISL_519566\|2020 | ···················· | ···················· | ···················· | ···················· | ···················· | ···················· |
| G | 3379 hCoV-19/Australia/VIC4239/2020\|EPI_ISL_519567\|2020 | ···················· | ···················· | ···················· | ···················· | ···················· | ···················· |
| G | 3380 hCoV-19/Australia/VIC4240/2020\|EPI_ISL_519568\|2020 | ···················· | ···················· | ···················· | ···················· | ···················· | ···················· |
| G | 3381 hCoV-19/Australia/VIC4241/2020\|EPI_ISL_519569\|2020 | ···················· | ···················· | ···················· | ···················· | ···················· | ···················· |
| G | 3382 hCoV-19/Australia/VIC4242/2020\|EPI_ISL_519570\|2020 | ···················· | ···················· | ···················· | ···················· | ···················· | ···················· |
| G | 3383 hCoV-19/Australia/VIC4243/2020\|EPI_ISL_519571\|2020 | ···················· | ···················· | ···················· | ···················· | ···················· | ···················· |
| G | 3384 hCoV-19/Australia/VIC4244/2020\|EPI_ISL_519572\|2020 | ···················· | ···················· | ···················· | ···················· | ···················· | ···················· |
| G | 3385 hCoV-19/Australia/VIC4245/2020\|EPI_ISL_519573\|2020 | ···················· | ···················· | ···················· | ···················· | ···················· | ···················· |
| G | 3386 hCoV-19/Australia/VIC4246/2020\|EPI_ISL_519574\|2020 | ···················· | ···················· | ···················· | ···················· | ···················· | ···················· |
| G | 3387 hCoV-19/Australia/VIC4247/2020\|EPI_ISL_519575\|2020 | ···················· | ···················· | ···················· | ···················· | ···················· | ···················· |
| G | 3388 hCoV-19/Australia/VIC4249/2020\|EPI_ISL_519577\|2020 | ···················· | ···················· | ···················· | ···················· | ···················· | ···················· |
| G | 3389 hCoV-19/Australia/VIC4250/2020\|EPI_ISL_519578\|2020 | ···················· | ···················· | ···················· | ···················· | ···················· | ···················· |
| G | 3390 hCoV-19/Australia/VIC4251/2020\|EPI_ISL_519579\|2020 | ···················· | ···················· | ···················· | ···················· | ···················· | ···················· |
| G | 3391 hCoV-19/Australia/VIC3805/2020\|EPI_ISL_520522\|2020 | ···················· | ···················· | ···················· | ···················· | ···················· | ···················· |
| G | 3392 hCoV-19/Australia/VIC3806/2020\|EPI_ISL_520523\|2020 | ···················· | ···················· | ···················· | ···················· | ···················· | ···················· |
| G | 3393 hCoV-19/Australia/VIC3807/2020\|EPI_ISL_520524\|2020 | ···················· | ···················· | ···················· | ···················· | ···················· | ···················· |
| G | 3394 hCoV-19/Australia/VIC3808/2020\|EPI_ISL_520525\|2020 | ···················· | ···················· | ···················· | ···················· | ···················· | ···················· |
| G | 3395 hCoV-19/Australia/VIC3812/2020\|EPI_ISL_520528\|2020 | ···················· | ···················· | ···················· | ···················· | ···················· | ···················· |
| G | 3396 hCoV-19/Australia/VIC3813/2020\|EPI_ISL_520529\|2020 | ···················· | ···················· | ···················· | ···················· | ···················· | ···················· |
| G | 3397 hCoV-19/Australia/VIC3814/2020\|EPI_ISL_520530\|2020 | ···················· | ···················· | ···················· | ···················· | ···················· | ···················· |
| G | 3398 hCoV-19/Australia/VIC3815/2020\|EPI_ISL_520531\|2020 | ···················· | ···················· | ···················· | ···················· | ···················· | ···················· |
| G | 3399 hCoV-19/Australia/VIC3816/2020\|EPI_ISL_520532\|2020 | ···················· | ···················· | ···················· | ···················· | ···················· | ···················· |
| G | 3400 hCoV-19/Australia/VIC3820/2020\|EPI_ISL_520533\|2020 | ···················· | ···················· | ···················· | ···················· | ···················· | ···················· |
| G | 3401 hCoV-19/Australia/VIC3821/2020\|EPI_ISL_520534\|2020 | ···················· | ···················· | ···················· | ···················· | ···················· | ···················· |
| G | 3402 hCoV-19/Australia/VIC3822/2020\|EPI_ISL_520535\|2020 | ···················· | ···················· | ···················· | ···················· | ···················· | ···················· |
| G | 3403 hCoV-19/Australia/VIC3823/2020\|EPI_ISL_520536\|2020 | ···················· | ···················· | ···················· | ···················· | ···················· | ···················· |
| G | 3404 hCoV-19/Australia/VIC3824/2020\|EPI_ISL_520537\|2020 | ···················· | ···················· | ···················· | ···················· | ···················· | ···················· |
| G | 3405 hCoV-19/Australia/VIC3825/2020\|EPI_ISL_520538\|2020 | ···················· | ···················· | ···················· | ···················· | ···················· | ···················· |
| G | 3406 hCoV-19/Australia/VIC3826/2020\|EPI_ISL_520539\|2020 | ···················· | ···················· | ···················· | ···················· | ···················· | ···················· |
| G | 3407 hCoV-19/Australia/VIC3827/2020\|EPI_ISL_520540\|2020 | ···················· | ···················· | ···················· | ···················· | ···················· | ···················· |
| G | 3408 hCoV-19/Australia/VIC3828/2020\|EPI_ISL_520541\|2020 | ···················· | ···················· | ···················· | ···················· | ···················· | ···················· |
| G | 3409 hCoV-19/Australia/VIC3829/2020\|EPI_ISL_520542\|2020 | ···················· | ···················· | ···················· | ···················· | ···················· | ···················· |
| G | 3410 hCoV-19/Australia/VIC3830/2020\|EPI_ISL_520543\|2020 | ···················· | ···················· | ···················· | ···················· | ···················· | ···················· |
| G | 3411 hCoV-19/Australia/VIC3831/2020\|EPI_ISL_520544\|2020 | ···················· | ···················· | ···················· | ···················· | ···················· | ···················· |
| G | 3412 hCoV-19/Australia/VIC3832/2020\|EPI_ISL_520545\|2020 | ···················· | ···················· | ···················· | ···················· | ···················· | ···················· |
| G | 3413 hCoV-19/Australia/VIC3833/2020\|EPI_ISL_520546\|2020 | ···················· | ···················· | ···················· | ···················· | ···················· | ···················· |
| G | 3414 hCoV-19/Australia/VIC3834/2020\|EPI_ISL_520547\|2020 | ···················· | ···················· | ···················· | ···················· | ···················· | ···················· |
| G | 3415 hCoV-19/Australia/VIC3835/2020\|EPI_ISL_520548\|2020 | ···················· | ···················· | ···················· | ···················· | ···················· | ···················· |
| G | 3416 hCoV-19/Australia/VIC3836/2020\|EPI_ISL_520549\|2020 | ···················· | ···················· | ···················· | ···················· | ···················· | ···················· |
| G | 3417 hCoV-19/Australia/VIC3837/2020\|EPI_ISL_520550\|2020 | ···················· | ···················· | ···················· | ···················· | ···················· | ···················· |
| G | 3419 hCoV-19/Australia/VIC3839/2020\|EPI_ISL_520552\|2020 | ···················· | ···················· | ···················· | ···················· | ···················· | ···················· |
| G | 3420 hCoV-19/Australia/VIC3841/2020\|EPI_ISL_520554\|2020 | ···················· | ···················· | ···················· | ···················· | ···················· | ···················· |
| G | 3421 hCoV-19/Australia/VIC3842/2020\|EPI_ISL_520555\|2020 | ···················· | ···················· | ···················· | ···················· | ···················· | ···················· |
| G | 3422 hCoV-19/Australia/VIC3843/2020\|EPI_ISL_520556\|2020 | ···················· | ···················· | ···················· | ···················· | ···················· | ···················· |
| G | 3423 hCoV-19/Australia/VIC3844/2020\|EPI_ISL_520557\|2020 | ···················· | ···················· | ···················· | ···················· | ···················· | ···················· |
| G | 3424 hCoV-19/Australia/VIC3845/2020\|EPI_ISL_520558\|2020 | ···················· | ···················· | ···················· | ···················· | ···················· | ···················· |
| G | 3425 hCoV-19/Australia/VIC3846/2020\|EPI_ISL_520559\|2020 | ···················· | ···················· | ···················· | ···················· | ···················· | ···················· |
| G | 3426 hCoV-19/Australia/VIC3847/2020\|EPI_ISL_520560\|2020 | ···················· | ···················· | ···················· | ···················· | ···················· | ···················· |
| G | 3427 hCoV-19/Australia/VIC3848/2020\|EPI_ISL_520561\|2020 | ···················· | ···················· | ···················· | ···················· | ···················· | ···················· |
| G | 3428 hCoV-19/Australia/VIC3849/2020\|EPI_ISL_520562\|2020 | ···················· | ···················· | ···················· | ···················· | ···················· | ···················· |
| G | 3429 hCoV-19/Australia/VIC3850/2020\|EPI_ISL_520563\|2020 | ···················· | ···················· | ···················· | ···················· | ···················· | ···················· |
| G | 3430 hCoV-19/Australia/VIC3851/2020\|EPI_ISL_520564\|2020 | ···················· | ···················· | ···················· | ···················· | ···················· | ···················· |
| G | 3431 hCoV-19/Australia/VIC3852/2020\|EPI_ISL_520565\|2020 | ···················· | ···················· | ···················· | ···················· | ···················· | ···················· |
| G | 3432 hCoV-19/Australia/VIC2970/2020\|EPI_ISL_521800\|2020 | ···················· | ···················· | ···················· | ···················· | ···················· | ···················· |
| G | 3433 hCoV-19/Australia/VIC2971/2020\|EPI_ISL_521801\|2020 | ···················· | ···················· | ···················· | ···················· | ···················· | ···················· |
| G | 3434 hCoV-19/Australia/VIC2972/2020\|EPI_ISL_521802\|2020 | ···················· | ···················· | ···················· | ···················· | ···················· | ···················· |
| G | 3435 hCoV-19/Australia/VIC2973/2020\|EPI_ISL_521803\|2020 | ···················· | ···················· | ···················· | ···················· | ···················· | ···················· |
| G | 3436 hCoV-19/Australia/VIC2974/2020\|EPI_ISL_521804\|2020 | ···················· | ···················· | ···················· | ···················· | ···················· | ···················· |
| G | 3437 hCoV-19/Australia/VIC2975/2020\|EPI_ISL_521805\|2020 | ···················· | ···················· | ···················· | ···················· | ···················· | ···················· |
| G | 3438 hCoV-19/Australia/VIC2976/2020\|EPI_ISL_521806\|2020 | ···················· | ···················· | ···················· | ···················· | ···················· | ···················· |
| G | 3439 hCoV-19/Australia/VIC2977/2020\|EPI_ISL_521807\|2020 | ···················· | ···················· | ···················· | ···················· | ···················· | ···················· |
| G | 3440 hCoV-19/Australia/VIC2978/2020\|EPI_ISL_521808\|2020 | ···················· | ···················· | ···················· | ···················· | ···················· | ···················· |
| G | 3441 hCoV-19/Australia/VIC2979/2020\|EPI_ISL_521809\|2020 | ···················· | ···················· | ···················· | ···················· | ···················· | ···················· |
| G | 3442 hCoV-19/Australia/VIC3024/2020\|EPI_ISL_521853\|2020 | ···················· | ···················· | ···················· | ···················· | ···················· | ···················· |
| G | 3443 hCoV-19/Australia/VIC3025/2020\|EPI_ISL_521854\|2020 | ···················· | ···················· | ···················· | ···················· | ···················· | ···················· |
| G | 3444 hCoV-19/Australia/VIC3027/2020\|EPI_ISL_521856\|2020 | ···················· | ···················· | ···················· | ···················· | ···················· | ···················· |
| G | 3445 hCoV-19/Australia/VIC3030/2020\|EPI_ISL_521859\|2020 | ···················· | ···················· | ···················· | ···················· | ···················· | ···················· |
| G | 3446 hCoV-19/Australia/NT51/2020\|EPI_ISL_521860\|2020-07 | ···················· | ···················· | ···················· | ···················· | ···················· | ···················· |
| G | 3448 hCoV-19/Australia/NT53/2020\|EPI_ISL_521862\|2020-06 | ···················· | ···················· | ···················· | ···················· | ···················· | ···················· |
| G | 3449 hCoV-19/Australia/VIC1386/2020\|EPI_ISL_521871\|2020 | ···················· | ···················· | ···················· | ···················· | ···················· | ···················· |
| G | 3450 hCoV-19/Australia/VIC1822/2020\|EPI_ISL_521872\|2020 | ···················· | ·A·················· | ···················· | ···················· | ···················· | ···A················ |
| G | 3451 hCoV-19/Australia/VIC1823/2020\|EPI_ISL_521873\|2020 | ···················· | ···················· | ···················· | ···················· | ···················· | ···················· |
| G | 3452 hCoV-19/Australia/VIC1824/2020\|EPI_ISL_521874\|2020 | ···················· | ·A·················· | ···················· | ···················· | ···················· | ···A················ |
| G | 3453 hCoV-19/Australia/VIC1825/2020\|EPI_ISL_521875\|2020 | ···················· | ···················· | ···················· | ···················· | ···················· | ···················· |
| G | 3454 hCoV-19/Australia/VIC1826/2020\|EPI_ISL_521876\|2020 | ···················· | ···················· | ···················· | ···················· | ···················· | ···················· |
| G | 3455 hCoV-19/Australia/VIC1828/2020\|EPI_ISL_521877\|2020 | ···················· | ···················· | ···················· | ···················· | ···················· | ···················· |
| G | 3456 hCoV-19/Australia/VIC1829/2020\|EPI_ISL_521878\|2020 | ···················· | ···················· | ···················· | ···················· | ···················· | ···················· |
| G | 3457 hCoV-19/Australia/VIC1831/2020\|EPI_ISL_521879\|2020 | ···················· | ···················· | ···················· | ···················· | ···················· | ···················· |
| G | 3458 hCoV-19/Australia/VIC1834/2020\|EPI_ISL_521880\|2020 | ···················· | ···················· | ···················· | ···················· | ···················· | ···················· |

| | | | | | | | |
|---|---|---|---|---|---|---|---|
| G | 3459 hCoV-19/Australia/VIC1835/2020\|EPI_ISL_521881\|2020 | ···················· | ···················· | ····················· | ···················· | ···················· | ···················· |
| G | 3460 hCoV-19/Australia/VIC1836/2020\|EPI_ISL_521882\|2020 | ···················· | ···················· | ····················· | ···················· | ···················· | ···················· |
| G | 3461 hCoV-19/Australia/VIC1837/2020\|EPI_ISL_521883\|2020 | ···················· | ···················· | ····················· | ···················· | ···················· | ···················· |
| G | 3462 hCoV-19/Australia/VIC1838/2020\|EPI_ISL_521884\|2020 | ···················· | ···················· | ····················· | ···················· | ···················· | ···················· |
| G | 3463 hCoV-19/Australia/VIC1845/2020\|EPI_ISL_521887\|2020 | ···················· | ···················· | ····················· | ···················· | ···················· | ···················· |
| G | 3464 hCoV-19/Australia/VIC1964/2020\|EPI_ISL_521892\|2020 | ···················· | ···················· | ····················· | ···················· | ···················· | ···················· |
| G | 3465 hCoV-19/Australia/VIC1970/2020\|EPI_ISL_521893\|2020 | ···················· | ···················· | ····················· | ···················· | ···················· | ···················· |
| G | 3466 hCoV-19/Australia/VIC1975/2020\|EPI_ISL_521894\|2020 | ···················· | ···················· | ····················· | ···················· | ···················· | ···················· |
| G | 3467 hCoV-19/Australia/VIC1984/2020\|EPI_ISL_521895\|2020 | ···················· | ···················· | ····················· | ···················· | ···················· | ···················· |
| G | 3468 hCoV-19/Russia/SPE-RII-15320V/2020\|EPI_ISL_524009\| | ···················· | ···················· | ····················· | ···················· | ···················· | ···················· |
| G | 3469 hCoV-19/Russia/SPE-RII-15911V/2020\|EPI_ISL_524010\| | ···················· | ···················· | ····················· | ···················· | ···················· | ···················· |
| G | 3470 hCoV-19/Russia/SPE-RII-16011V/2020\|EPI_ISL_524011\| | ···················· | ···················· | ····················· | ···················· | ···················· | ···················· |
| G | 3471 hCoV-19/Russia/SPE-RII-16850V/2020\|EPI_ISL_524012\| | ···················· | ···················· | ····················· | ···················· | ···················· | ···················· |
| G | 3472 hCoV-19/Russia/SPE-RII-17217V/2020\|EPI_ISL_524013\| | ···················· | ···················· | ····················· | ···················· | ···················· | ···················· |
| G | 3473 hCoV-19/Russia/SPE-RII-17291V/2020\|EPI_ISL_524014\| | ···················· | ···················· | ····················· | ···················· | ···················· | ···················· |
| G | 3474 hCoV-19/Russia/SPE-RII-17684V/2020\|EPI_ISL_524015\| | ···················· | ···················· | ····················· | ···················· | ···················· | ···················· |
| G | 3475 hCoV-19/Russia/SPE-RII-17942V/2020\|EPI_ISL_524016\| | ···················· | ···················· | ····················· | ···················· | ···················· | ···················· |
| G | 3476 hCoV-19/Russia/SPE-RII-18220V/2020\|EPI_ISL_524017\| | ···················· | ···················· | ····················· | ···················· | ···················· | ···················· |
| G | 3477 hCoV-19/Russia/SPE-RII-18357V/2020\|EPI_ISL_524018\| | ···················· | ···················· | ····················· | ···················· | ···················· | ···················· |
| G | 3478 hCoV-19/Russia/SPE-RII-18358V/2020\|EPI_ISL_524019\| | ···················· | ···················· | ····················· | ···················· | ···················· | ···················· |
| G | 3479 hCoV-19/Russia/SPE-RII-18574V/2020\|EPI_ISL_524020\| | ···················· | ···················· | ····················· | ···················· | ···················· | ···················· |
| G | 3480 hCoV-19/Russia/SPE-RII-18663V/2020\|EPI_ISL_524021\| | ···················· | ···················· | ····················· | ···················· | ···················· | ···················· |
| G | 3481 hCoV-19/Russia/SPE-RII-18966V/2020\|EPI_ISL_524022\| | ···················· | ···················· | ····················· | ···················· | ···················· | ···················· |
| G | 3482 hCoV-19/Russia/SPE-RII-19109V/2020\|EPI_ISL_524023\| | ···················· | ···················· | ····················· | ···················· | ···················· | ···················· |
| G | 3483 hCoV-19/Russia/SPE-RII-19422V/2020\|EPI_ISL_524024\| | ···················· | ···················· | ····················· | ···················· | ···················· | ···················· |
| G | 3484 hCoV-19/Russia/SPE-RII-19426V/2020\|EPI_ISL_524025\| | ···················· | ···················· | ····················· | ···················· | ···················· | ···················· |
| G | 3485 hCoV-19/Russia/SPE-RII-19578V/2020\|EPI_ISL_524026\| | ···················· | ···················· | ····················· | ···················· | ···················· | ···················· |
| G | 3486 hCoV-19/Russia/SPE-RII-19602V/2020\|EPI_ISL_524027\| | ···················· | ···················· | ····················· | ···················· | ···················· | ···················· |
| G | 3487 hCoV-19/Russia/SPE-RII-19652V/2020\|EPI_ISL_524028\| | ···················· | ···················· | ····················· | ···················· | ···················· | ···················· |
| G | 3488 hCoV-19/Russia/SPE-RII-20404V/2020\|EPI_ISL_524029\| | ···················· | ···················· | ····················· | ···················· | ···················· | ···················· |
| G | 3489 hCoV-19/Russia/SPE-RII-20906V/2020\|EPI_ISL_524030\| | ···················· | ···················· | ····················· | ···················· | ···················· | ···················· |
| G | 3490 hCoV-19/Russia/SPE-RII-21056V/2020\|EPI_ISL_524031\| | ···················· | ···················· | ····················· | ···················· | ···················· | ···················· |
| G | 3491 hCoV-19/Russia/SPE-RII-21119V/2020\|EPI_ISL_524032\| | ···················· | ···················· | ····················· | ···················· | ···················· | ···················· |
| G | 3492 hCoV-19/Russia/SPE-RII-21357V/2020\|EPI_ISL_524033\| | ···················· | ···················· | ····················· | ···················· | ···················· | ···················· |
| G | 3493 hCoV-19/Russia/SPE-RII-21358V/2020\|EPI_ISL_524034\| | ···················· | ···················· | ····················· | ···················· | ···················· | ···················· |
| G | 3494 hCoV-19/Russia/SPE-RII-22043V/2020\|EPI_ISL_524035\| | ···················· | ···················· | ····················· | ···················· | ···················· | ···················· |
| G | 3495 hCoV-19/Russia/SPE-RII-22068V/2020\|EPI_ISL_524036\| | ···················· | ···················· | ····················· | ···················· | ···················· | ···················· |
| G | 3496 hCoV-19/Russia/SPE-RII-22069V/2020\|EPI_ISL_524037\| | ···················· | ···················· | ····················· | ···················· | ···················· | ···················· |
| G | 3497 hCoV-19/Russia/SPE-RII-22086V/2020\|EPI_ISL_524038\| | ···················· | ···················· | ····················· | ···················· | ···················· | ···················· |
| G | 3498 hCoV-19/Russia/SPE-RII-22207V/2020\|EPI_ISL_524039\| | ···················· | ···················· | ····················· | ···················· | ···················· | ···················· |
| G | 3499 hCoV-19/Russia/SPE-RII-22266V/2020\|EPI_ISL_524040\| | ···················· | ···················· | ····················· | ···················· | ···················· | ···················· |
| G | 3500 hCoV-19/Russia/SPE-RII-22269V/2020\|EPI_ISL_524041\| | ···················· | ···················· | ····················· | ···················· | ···················· | ···················· |
| G | 3501 hCoV-19/Russia/SPE-RII-22276V/2020\|EPI_ISL_524042\| | ···················· | ···················· | ····················· | ···················· | ···················· | ···················· |
| G | 3502 hCoV-19/Russia/SPE-RII-22311V/2020\|EPI_ISL_524043\| | ···················· | ···················· | ····················· | ···················· | ···················· | ···················· |
| G | 3503 hCoV-19/Russia/SPE-RII-22388V/2020\|EPI_ISL_524044\| | ···················· | ···················· | ····················· | ···················· | ···················· | ···················· |
| G | 3504 hCoV-19/Russia/SPE-RII-22478V/2020\|EPI_ISL_524045\| | ···················· | ···················· | ····················· | ···················· | ···················· | ···················· |
| G | 3505 hCoV-19/Russia/SPE-RII-23386V/2020\|EPI_ISL_524046\| | ···················· | ···················· | ····················· | ···················· | ···················· | ···················· |
| G | 3506 hCoV-19/Russia/SPE-RII-24810V/2020\|EPI_ISL_524047\| | ···················· | ···················· | ····················· | ···················· | ···················· | ···················· |
| G | 3507 hCoV-19/Russia/SPE-RII-6425V/2020\|EPI_ISL_524048\|2 | ···················· | ···················· | ····················· | ···················· | ···················· | ···················· |
| G | 3508 hCoV-19/Russia/SPE-RII-7388V/2020\|EPI_ISL_524049\|2 | ···················· | ···················· | ····················· | ···················· | ···················· | ···················· |
| G | 3509 hCoV-19/Russia/SPE-RII-8532V/2020\|EPI_ISL_524050\|2 | ···················· | ···················· | ····················· | ···················· | ···················· | ···················· |
| G | 3510 hCoV-19/Russia/SPE-RII-8542V/2020\|EPI_ISL_524051\|2 | ···················· | ···················· | ····················· | ···················· | ···················· | ···················· |
| G | 3511 hCoV-19/Russia/SPE-RII-9178V/2020\|EPI_ISL_524052\|2 | ···················· | ···················· | ····················· | ···················· | ···················· | ···················· |
| G | 3512 hCoV-19/USA/UT-08268/2020\|EPI_ISL_524074\|2020-05-1 | ···················· | ···················· | ····················· | ···················· | ···················· | ···················· |
| G | 3513 hCoV-19/USA/UT-08269/2020\|EPI_ISL_524075\|2020-05-2 | ···················· | ···················· | ····················· | ···················· | ···················· | ···················· |
| G | 3514 hCoV-19/USA/UT-08272/2020\|EPI_ISL_524076\|2020-05-2 | ···················· | ···················· | ····················· | ···················· | ···················· | ···················· |
| G | 3515 hCoV-19/USA/UT-08275/2020\|EPI_ISL_524077\|2020-05-2 | ···················· | ·A·················· | ····················· | ···················· | ···················· | ···················· |
| G | 3516 hCoV-19/USA/UT-08276/2020\|EPI_ISL_524078\|2020-05-2 | ···················· | ···················· | ····················· | ···················· | ···················· | ···················· |
| G | 3517 hCoV-19/USA/UT-08279/2020\|EPI_ISL_524080\|2020-05-1 | ···················· | ···················· | ····················· | ···················· | ···················· | ···················· |
| G | 3518 hCoV-19/USA/UT-08281/2020\|EPI_ISL_524081\|2020-05-2 | ···················· | ·A·················· | ····················· | ···················· | ···················· | ···················· |
| G | 3519 hCoV-19/USA/UT-08282/2020\|EPI_ISL_524082\|2020-05-1 | ···················· | ···················· | ····················· | ···················· | ···················· | ···················· |
| G | 3520 hCoV-19/USA/UT-08285/2020\|EPI_ISL_524084\|2020-05-2 | ···················· | ···················· | ····················· | ···················· | ···················· | ···················· |
| G | 3521 hCoV-19/USA/UT-08288/2020\|EPI_ISL_524087\|2020-05-2 | ···················· | ···················· | ····················· | ···················· | ···················· | ···················· |
| G | 3522 hCoV-19/USA/UT-08290/2020\|EPI_ISL_524089\|2020-05-1 | ···················· | ···················· | ····················· | ···················· | ···················· | ···················· |
| G | 3523 hCoV-19/USA/UT-UPHL-04473/2020\|EPI_ISL_525340\|2020 | ···················· | ···················· | ····················· | ···················· | ···················· | ···················· |
| G | 3524 hCoV-19/USA/UT-UPHL-04478/2020\|EPI_ISL_525343\|2020 | ···················· | ·A·················· | ····················· | ···················· | ···················· | ···················· |
| G | 3525 hCoV-19/USA/UT-UPHL-04483/2020\|EPI_ISL_525344\|2020 | ···················· | ···················· | ····················· | ···················· | ···················· | ···················· |
| G | 3526 hCoV-19/USA/UT-UPHL-04485/2020\|EPI_ISL_525345\|2020 | ···················· | ···················· | ····················· | ···················· | ···················· | ···················· |
| G | 3527 hCoV-19/USA/UT-UPHL-04487/2020\|EPI_ISL_525346\|2020 | ···················· | ···················· | ····················· | ···················· | ···················· | ···················· |
| G | 3528 hCoV-19/USA/UT-UPHL-08760/2020\|EPI_ISL_525347\|2020 | ···················· | ·A·················· | ····················· | ···················· | ···················· | ···················· |
| G | 3529 hCoV-19/USA/UT-UPHL-08762/2020\|EPI_ISL_525348\|2020 | ···················· | ·A·················· | ····················· | ···················· | ···················· | ···················· |
| G | 3530 hCoV-19/USA/UT-UPHL-08764/2020\|EPI_ISL_525350\|2020 | ···················· | ·A·················· | ····················· | ···················· | ···················· | ···················· |
| G | 3531 hCoV-19/USA/UT-UPHL-08765/2020\|EPI_ISL_525351\|2020 | ···················· | ·A·················· | ····················· | ···················· | ···················· | ···················· |
| G | 3532 hCoV-19/Ireland/LS-NVRL-71IRL93650/2020\|EPI_ISL_52 | ···················· | ···················· | ····················· | ···················· | ···················· | ···················· |
| G | 3533 hCoV-19/Ireland/LS-NVRL-71IRL93658/2020\|EPI_ISL_52 | ···················· | ···················· | ····················· | ···················· | ···················· | ···················· |
| G | 3534 hCoV-19/Ireland/KE-NVRL-71IRL94136/2020\|EPI_ISL_52 | ···················· | ···················· | ····················· | ···················· | ···················· | ···················· |
| G | 3535 hCoV-19/Ireland/KE-NVRL-71IRL94145/2020\|EPI_ISL_52 | ···················· | ···················· | ····················· | ···················· | ···················· | ···················· |
| G | 3536 hCoV-19/Ireland/D-NVRL-71IRL94252/2020\|EPI_ISL_525 | ···················· | ···················· | ····················· | ···················· | ···················· | ···················· |
| G | 3537 hCoV-19/Ireland/D-NVRL-71IRL94359/2020\|EPI_ISL_525 | ···················· | ···················· | ····················· | ···················· | ···················· | ···················· |
| G | 3538 hCoV-19/Ireland/OY-NVRL-71IRL94548/2020\|EPI_ISL_52 | ···················· | ···················· | ····················· | ···················· | ···················· | ···················· |

| | | | | | | | |
|---|---|---|---|---|---|---|---|
| G | 3539 hCoV-19/Ireland/MH-NVRL-71IRL94783/2020\|EPI_ISL_52 | ···················· | ···················· | ···················· | ···················· | ···················· | ···················· |
| G | 3540 hCoV-19/Ireland/D-NVRL-71IRL94844/2020\|EPI_ISL_525 | ···················· | ···················· | ···················· | ···················· | ···················· | ···················· |
| G | 3541 hCoV-19/Ireland/LH-NVRL-71IRL95259/2020\|EPI_ISL_52 | ···················· | ···················· | ···················· | ···················· | ···················· | ···················· |
| G | 3542 hCoV-19/Ireland/MH-NVRL-71IRL95385/2020\|EPI_ISL_52 | ···················· | ···················· | ···················· | ···················· | ···················· | ···················· |
| G | 3543 hCoV-19/Ireland/D-NVRL-71IRL95471/2020\|EPI_ISL_525 | ···················· | ···················· | ···················· | ···················· | ···················· | ···················· |
| G | 3544 hCoV-19/Ireland/D-NVRL-71IRL95850/2020\|EPI_ISL_525 | ···················· | ···················· | ···················· | ···················· | ···················· | ···················· |
| G | 3545 hCoV-19/Ireland/D-NVRL-71IRL95870/2020\|EPI_ISL_525 | ···················· | ···················· | ···················· | ···················· | ···················· | ···················· |
| G | 3546 hCoV-19/Ireland/WX-NVRL-71IRL95947/2020\|EPI_ISL_52 | ···················· | ···················· | ···················· | ···················· | ···················· | ···················· |
| G | 3547 hCoV-19/Ireland/WX-NVRL-71IRL95961/2020\|EPI_ISL_52 | ···················· | ···················· | ···················· | ···················· | ···················· | ···················· |
| G | 3548 hCoV-19/Ireland/CN-NVRL-71IRL96106/2020\|EPI_ISL_52 | ···················· | ···················· | ···················· | ···················· | ···················· | ···················· |
| G | 3549 hCoV-19/Ireland/D-NVRL-71IRL96127/2020\|EPI_ISL_525 | ···················· | ···················· | ···················· | ···················· | ···················· | ···················· |
| G | 3550 hCoV-19/Ireland/D-NVRL-71IRL96183/2020\|EPI_ISL_525 | ···················· | ···················· | ···················· | ···················· | ···················· | ···················· |
| G | 3552 hCoV-19/Ireland/KE-NVRL-71IRL96304/2020\|EPI_ISL_52 | ···················· | ···················· | ···················· | ···················· | ···················· | ···················· |
| G | 3553 hCoV-19/Ireland/LS-NVRL-71IRL96316/2020\|EPI_ISL_52 | ···················· | ···················· | ···················· | ···················· | ···················· | ···················· |
| G | 3554 hCoV-19/Ireland/KE-NVRL-71IRL96353/2020\|EPI_ISL_52 | ···················· | ···················· | ···················· | ···················· | ···················· | ···················· |
| G | 3555 hCoV-19/Ireland/KE-NVRL-71IRL96356/2020\|EPI_ISL_52 | ···················· | ···················· | ···················· | ···················· | ···················· | ···················· |
| G | 3556 hCoV-19/Ireland/KE-NVRL-71IRL96368/2020\|EPI_ISL_52 | ···················· | ···················· | ···················· | ···················· | ···················· | ···················· |
| G | 3557 hCoV-19/Ireland/KE-NVRL-71IRL96371/2020\|EPI_ISL_52 | ···················· | ···················· | ···················· | ···················· | ···················· | ···················· |
| G | 3558 hCoV-19/Ireland/LS-NVRL-71IRL96375/2020\|EPI_ISL_52 | ···················· | ···················· | ···················· | ···················· | ···················· | ···················· |
| G | 3559 hCoV-19/Ireland/KE-NVRL-71IRL96376/2020\|EPI_ISL_52 | ···················· | ···················· | ···················· | ···················· | ···················· | ···················· |
| G | 3560 hCoV-19/Ireland/KE-NVRL-71IRL96380/2020\|EPI_ISL_52 | ···················· | ···················· | ···················· | ···················· | ···················· | ···················· |
| G | 3561 hCoV-19/Ireland/KE-NVRL-71IRL97553/2020\|EPI_ISL_52 | ···················· | ···················· | ···················· | ···················· | ···················· | ···················· |
| G | 3562 hCoV-19/Ireland/KE-NVRL-71IRL98494/2020\|EPI_ISL_52 | ···················· | ···················· | ···················· | ···················· | ···················· | ···················· |
| G | 3563 hCoV-19/Ireland/KE-NVRL-71IRL98572/2020\|EPI_ISL_52 | ···················· | ···················· | ···················· | ···················· | ···················· | ···················· |
| G | 3564 hCoV-19/Ireland/LS-NVRL-71IRL98729/2020\|EPI_ISL_52 | ···················· | ···················· | ···················· | ···················· | ···················· | ···················· |
| G | 3565 hCoV-19/Ireland/OY-NVRL-72IRL00272/2020\|EPI_ISL_52 | ···················· | ···················· | ···················· | ···················· | ···················· | ···················· |
| G | 3566 hCoV-19/Ireland/OY-NVRL-72IRL00274/2020\|EPI_ISL_52 | ···················· | ···················· | ···················· | ···················· | ···················· | ···················· |
| G | 3567 hCoV-19/USA/FL-BPHL-1184/2020\|EPI_ISL_526670\|2020- | ···················· | ···················· | ···················· | ···················· | ···················· | ···················· |
| G | 3568 hCoV-19/USA/FL-BPHL-1185/2020\|EPI_ISL_526671\|2020- | ···················· | ···················· | ···················· | ···················· | ···················· | ···················· |
| G | 3569 hCoV-19/USA/FL-BPHL-1186/2020\|EPI_ISL_526672\|2020- | ···················· | ···················· | ···················· | ···················· | ···················· | ···················· |
| G | 3570 hCoV-19/USA/FL-BPHL-1187/2020\|EPI_ISL_526673\|2020- | ···················· | ···················· | ···················· | ···················· | ···················· | ···················· |
| G | 3571 hCoV-19/USA/FL-BPHL-1188/2020\|EPI_ISL_526674\|2020- | ···················· | ···················· | ···················· | ···················· | ···················· | ···················· |
| G | 3572 hCoV-19/USA/FL-BPHL-1189/2020\|EPI_ISL_526675\|2020- | ···················· | ·A·················· | ···················· | ···················· | ···················· | ···················· |
| G | 3574 hCoV-19/USA/FL-BPHL-1191/2020\|EPI_ISL_526677\|2020- | ···················· | ·A·················· | ···················· | ···················· | ···················· | ···················· |
| G | 3575 hCoV-19/USA/FL-BPHL-1192/2020\|EPI_ISL_526678\|2020- | ···················· | ···················· | ···················· | ···················· | ···················· | ···················· |
| G | 3576 hCoV-19/USA/FL-BPHL-1193/2020\|EPI_ISL_526679\|2020- | ···················· | ···················· | ···················· | ···················· | ···················· | ···················· |
| G | 3577 hCoV-19/USA/FL-BPHL-1194/2020\|EPI_ISL_526680\|2020- | ···················· | ···················· | ···················· | ···················· | ···················· | ···················· |
| G | 3578 hCoV-19/USA/FL-BPHL-1195/2020\|EPI_ISL_526681\|2020- | ···················· | ···················· | ···················· | ···················· | ···················· | ···················· |
| G | 3579 hCoV-19/USA/FL-BPHL-1196/2020\|EPI_ISL_526682\|2020- | ···················· | ···················· | ···················· | ···················· | ···················· | ···················· |
| G | 3580 hCoV-19/USA/NY-MH001867/2020\|EPI_ISL_526686\|2020-0 | ···················· | ···················· | ···················· | ···················· | ···················· | ···················· |
| G | 3581 hCoV-19/USA/NY-MH001827/2020\|EPI_ISL_526687\|2020-0 | ···················· | ···················· | ···················· | ···················· | ···················· | ···················· |
| G | 3582 hCoV-19/USA/NY-MH001832/2020\|EPI_ISL_526688\|2020-0 | ···················· | ···················· | ···················· | ···················· | ···················· | ···················· |
| G | 3583 hCoV-19/Australia/NSW772/2020\|EPI_ISL_526689\|2020- | ···················· | ···················· | ···················· | ···················· | ···················· | ···················· |
| G | 3584 hCoV-19/Australia/NSW873/2020\|EPI_ISL_526697\|2020- | ···················· | ···················· | ···················· | ···················· | ···················· | ···················· |
| G | 3585 hCoV-19/South Africa/KRISP-K002444/2020\|EPI_ISL_51 | ···················· | ···················· | ···················· | ···················· | ···················· | ···················· |
| G | 3586 hCoV-19/South Africa/KRISP-K002445/2020\|EPI_ISL_51 | ···················· | ···················· | ···················· | ···················· | ···················· | ···················· |
| G | 3587 hCoV-19/South Africa/KRISP-K002448/2020\|EPI_ISL_51 | ···················· | ···················· | ···················· | ···················· | ···················· | ···················· |
| G | 3588 hCoV-19/South Africa/KRISP-K002449/2020\|EPI_ISL_51 | ···················· | ···················· | ···················· | ···················· | ···················· | ···················· |
| G | 3589 hCoV-19/South Africa/KRISP-K002451/2020\|EPI_ISL_51 | ···················· | ···················· | ···················· | ···················· | ···················· | ···················· |
| G | 3590 hCoV-19/South Africa/KRISP-K002454/2020\|EPI_ISL_51 | ···················· | ···················· | ···················· | ···················· | ···················· | ···················· |
| G | 3591 hCoV-19/South Africa/KRISP-K002455/2020\|EPI_ISL_51 | ···················· | ···················· | ···················· | ···················· | ···················· | ···················· |
| G | 3592 hCoV-19/South Africa/KRISP-K002456/2020\|EPI_ISL_51 | ···················· | ···················· | ···················· | ···················· | ···················· | ···················· |
| G | 3593 hCoV-19/South Africa/KRISP-K002457/2020\|EPI_ISL_51 | ···················· | ···················· | ···················· | ···················· | ···················· | ···················· |
| G | 3594 hCoV-19/South Africa/KRISP-K002460/2020\|EPI_ISL_51 | ···················· | ···················· | ···················· | ···················· | ···················· | ···················· |
| G | 3595 hCoV-19/South Africa/KRISP-K002461/2020\|EPI_ISL_51 | ················G··· | ···················· | ···················· | ···················· | ···················· | ···················· |
| G | 3596 hCoV-19/South Africa/KRISP-K002463/2020\|EPI_ISL_51 | ···················· | ···················· | ···················· | ···················· | ···················· | ···················· |
| G | 3597 hCoV-19/South Africa/KRISP-K002464/2020\|EPI_ISL_51 | ···················· | ···················· | ···················· | ···················· | ···················· | ···················· |
| G | 3598 hCoV-19/South Africa/KRISP-K002465/2020\|EPI_ISL_51 | ···················· | ···················· | ···················· | ···················· | ···················· | ···················· |
| G | 3599 hCoV-19/South Africa/KRISP-K002466/2020\|EPI_ISL_51 | ···················· | ···················· | ···················· | ···················· | ···················· | ···················· |
| G | 3600 hCoV-19/South Africa/KRISP-K002467/2020\|EPI_ISL_51 | ···················· | ···················· | ···················· | ···················· | ···················· | ···················· |
| G | 3601 hCoV-19/South Africa/KRISP-K002472/2020\|EPI_ISL_51 | ···················· | ···················· | ···················· | ···················· | ···················· | ···················· |
| G | 3602 hCoV-19/South Africa/KRISP-K002474/2020\|EPI_ISL_51 | ···················· | ···················· | ···················· | ···················· | ···················· | ···················· |
| G | 3603 hCoV-19/Australia/VIC6022/2020\|EPI_ISL_518063\|2020 | ···················· | ···················· | ···················· | ···················· | ···················· | ···················· |
| G | 3604 hCoV-19/Australia/VIC6023/2020\|EPI_ISL_518064\|2020 | ···················· | ···················· | ···················· | ···················· | ···················· | ···················· |
| G | 3605 hCoV-19/Australia/VIC6024/2020\|EPI_ISL_518065\|2020 | ···················· | ···················· | ···················· | ···················· | ···················· | ···················· |
| G | 3606 hCoV-19/Australia/VIC6025/2020\|EPI_ISL_518066\|2020 | ···················· | ···················· | ···················· | ···················· | ···················· | ···················· |
| G | 3607 hCoV-19/Australia/VIC6026/2020\|EPI_ISL_518067\|2020 | ···················· | ···················· | ···················· | ···················· | ···················· | ···················· |
| G | 3608 hCoV-19/Australia/VIC6027/2020\|EPI_ISL_518068\|2020 | ···················· | ···················· | ···················· | ···················· | ···················· | ···················· |
| G | 3609 hCoV-19/Australia/VIC6028/2020\|EPI_ISL_518069\|2020 | ···················· | ···················· | ···················· | ···················· | ···················· | ···················· |
| G | 3610 hCoV-19/Australia/VIC6029/2020\|EPI_ISL_518070\|2020 | ···················· | ···················· | ···················· | ···················· | ···················· | ···················· |
| G | 3611 hCoV-19/Australia/VIC6030/2020\|EPI_ISL_518071\|2020 | ···················· | ···················· | ···················· | ···················· | ···················· | ···················· |
| G | 3612 hCoV-19/Australia/VIC6031/2020\|EPI_ISL_518072\|2020 | ···················· | ···················· | ···················· | ···················· | ···················· | ···················· |
| G | 3613 hCoV-19/Australia/VIC6032/2020\|EPI_ISL_518073\|2020 | ···················· | ···················· | ···················· | ···················· | ···················· | ···················· |
| G | 3614 hCoV-19/Australia/VIC6033/2020\|EPI_ISL_518074\|2020 | ···················· | ···················· | ···················· | ···················· | ···················· | ···················· |
| G | 3615 hCoV-19/Australia/VIC6034/2020\|EPI_ISL_518075\|2020 | ···················· | ···················· | ···················· | ···················· | ···················· | ···················· |
| G | 3616 hCoV-19/Australia/VIC6035/2020\|EPI_ISL_518076\|2020 | ···················· | ···················· | ···················· | ···················· | ···················· | ···················· |
| G | 3617 hCoV-19/Australia/VIC6036/2020\|EPI_ISL_518077\|2020 | ···················· | ···················· | ···················· | ···················· | ···················· | ···················· |
| G | 3618 hCoV-19/Australia/VIC6037/2020\|EPI_ISL_518078\|2020 | ···················· | ···················· | ···················· | ···················· | ···················· | ···················· |
| G | 3619 hCoV-19/Australia/VIC6038/2020\|EPI_ISL_518079\|2020 | ···················· | ···················· | ···················· | ···················· | ···················· | ···················· |
| G | 3620 hCoV-19/Australia/VIC6039/2020\|EPI_ISL_518080\|2020 | ···················· | ···················· | ···················· | ···················· | ···················· | ···················· |

| | | | | | | | | |
|---|---|---|---|---|---|---|---|---|
| G | 3621 | hCoV-19/Australia/VIC6040/2020\|EPI_ISL_518081\|2020 | .................... | .................... | ..................... | .................... | .................... | .................... |
| G | 3622 | hCoV-19/Australia/VIC6041/2020\|EPI_ISL_518082\|2020 | .................... | .................... | ..................... | .................... | .................... | .................... |
| G | 3623 | hCoV-19/Australia/VIC6042/2020\|EPI_ISL_518083\|2020 | .................... | .................... | ..................... | .................... | .................... | .................... |
| G | 3624 | hCoV-19/Australia/VIC6043/2020\|EPI_ISL_518084\|2020 | .................... | .................... | ..................... | .................... | .................... | .................... |
| G | 3625 | hCoV-19/Australia/VIC6045/2020\|EPI_ISL_518086\|2020 | .................... | .................... | ..................... | .................... | .................... | .................... |
| G | 3626 | hCoV-19/Australia/VIC6048/2020\|EPI_ISL_518088\|2020 | .................... | .................... | ..................... | .................... | .................... | .................... |
| G | 3627 | hCoV-19/Australia/VIC6049/2020\|EPI_ISL_518089\|2020 | .................... | .................... | ..................... | .................... | .................... | .................... |
| G | 3628 | hCoV-19/Australia/VIC6198/2020\|EPI_ISL_518200\|2020 | .................... | .................... | ..................... | .................... | .................... | .................... |
| G | 3629 | hCoV-19/Australia/VIC6199/2020\|EPI_ISL_518201\|2020 | .................... | .................... | ..................... | .................... | .................... | .................... |
| G | 3630 | hCoV-19/Australia/VIC6200/2020\|EPI_ISL_518202\|2020 | .................... | .................... | ..................... | .................... | .................... | .................... |
| G | 3631 | hCoV-19/Australia/VIC6201/2020\|EPI_ISL_518203\|2020 | .................... | .................... | ..................... | .................... | .................... | .................... |
| G | 3632 | hCoV-19/Australia/VIC6203/2020\|EPI_ISL_518204\|2020 | .................... | .................... | ..................... | .................... | .................... | .................... |
| G | 3633 | hCoV-19/Australia/VIC6204/2020\|EPI_ISL_518205\|2020 | .................... | .................... | ..................... | .................... | .................... | .................... |
| G | 3634 | hCoV-19/Australia/VIC6205/2020\|EPI_ISL_518206\|2020 | .................... | .................... | ..................... | .................... | .................... | .................... |
| G | 3635 | hCoV-19/Australia/VIC6206/2020\|EPI_ISL_518207\|2020 | .................... | .................... | ..................... | .................... | .................... | .................... |
| G | 3636 | hCoV-19/Australia/VIC6207/2020\|EPI_ISL_518208\|2020 | .................... | .................... | ..................... | .................... | .................... | .................... |
| G | 3637 | hCoV-19/Australia/VIC6208/2020\|EPI_ISL_518209\|2020 | .................... | .................... | ..................... | .................... | .................... | .................... |
| G | 3638 | hCoV-19/Australia/VIC6217/2020\|EPI_ISL_518215\|2020 | .................... | .................... | ..................... | .................... | .................... | .................... |
| G | 3639 | hCoV-19/Australia/VIC6219/2020\|EPI_ISL_518217\|2020 | .................... | .................... | ..................... | .................... | .................... | .................... |
| G | 3640 | hCoV-19/Australia/VIC6220/2020\|EPI_ISL_518218\|2020 | .................... | .................... | ..................... | .................... | .................... | .................... |
| G | 3641 | hCoV-19/Australia/VIC6221/2020\|EPI_ISL_518219\|2020 | .................... | .................... | ..................... | .................... | .................... | .................... |
| G | 3642 | hCoV-19/Australia/VIC3956/2020\|EPI_ISL_519350\|2020 | .................... | .................... | ..................... | .................... | .................... | .................... |
| G | 3643 | hCoV-19/Australia/VIC3968/2020\|EPI_ISL_519360\|2020 | .................... | .................... | ..................... | .................... | .................... | .................... |
| G | 3644 | hCoV-19/Australia/VIC3969/2020\|EPI_ISL_519361\|2020 | .................... | .................... | ..................... | .................... | .................... | .................... |
| G | 3645 | hCoV-19/Australia/VIC3970/2020\|EPI_ISL_519362\|2020 | .................... | .................... | ..................... | .................... | .................... | .................... |
| G | 3646 | hCoV-19/Australia/VIC3971/2020\|EPI_ISL_519363\|2020 | .................... | .................... | ..................... | .................... | .................... | .................... |
| G | 3647 | hCoV-19/Australia/VIC3972/2020\|EPI_ISL_519364\|2020 | .................... | .................... | ..................... | .................... | .................... | .................... |
| G | 3648 | hCoV-19/Australia/VIC3973/2020\|EPI_ISL_519365\|2020 | .................... | .................... | ..................... | .................... | .................... | .................... |
| G | 3649 | hCoV-19/Australia/VIC3974/2020\|EPI_ISL_519366\|2020 | .................... | .................... | ..................... | .................... | .................... | .................... |
| G | 3650 | hCoV-19/Australia/VIC3975/2020\|EPI_ISL_519367\|2020 | .................... | .................... | ..................... | .................... | .................... | .................... |
| G | 3651 | hCoV-19/Australia/VIC3976/2020\|EPI_ISL_519368\|2020 | .................... | .................... | ..................... | .................... | .................... | .................... |
| G | 3652 | hCoV-19/Australia/VIC3977/2020\|EPI_ISL_519369\|2020 | .................... | .................... | ..................... | .................... | .................... | .................... |
| G | 3653 | hCoV-19/Australia/VIC3979/2020\|EPI_ISL_519371\|2020 | .................... | .................... | ..................... | .................... | .................... | .................... |
| G | 3654 | hCoV-19/Australia/VIC3980/2020\|EPI_ISL_519372\|2020 | .................... | .................... | ..................... | .................... | .................... | .................... |
| G | 3655 | hCoV-19/Australia/VIC3981/2020\|EPI_ISL_519373\|2020 | .................... | .................... | ..................... | .................... | .................... | .................... |
| G | 3656 | hCoV-19/Australia/VIC3982/2020\|EPI_ISL_519374\|2020 | .................... | .................... | ..................... | .................... | .................... | .................... |
| G | 3657 | hCoV-19/Australia/VIC3985/2020\|EPI_ISL_519375\|2020 | .................... | .................... | ..................... | .................... | .................... | .................... |
| G | 3658 | hCoV-19/Australia/VIC3986/2020\|EPI_ISL_519376\|2020 | .................... | .................... | ..................... | .................... | .................... | .................... |
| G | 3659 | hCoV-19/Australia/VIC3987/2020\|EPI_ISL_519377\|2020 | .................... | .................... | ..................... | .................... | .................... | .................... |
| G | 3660 | hCoV-19/Australia/VIC3988/2020\|EPI_ISL_519378\|2020 | .................... | .................... | ..................... | .................... | .................... | .................... |
| G | 3661 | hCoV-19/Australia/VIC3989/2020\|EPI_ISL_519379\|2020 | .................... | .................... | ..................... | .................... | .................... | .................... |
| G | 3662 | hCoV-19/Australia/VIC3990/2020\|EPI_ISL_519380\|2020 | .................... | .................... | ..................... | .................... | .................... | .................... |
| G | 3663 | hCoV-19/Australia/VIC3991/2020\|EPI_ISL_519381\|2020 | .................... | .................... | ..................... | .................... | .................... | .................... |
| G | 3664 | hCoV-19/Australia/VIC3992/2020\|EPI_ISL_519382\|2020 | .................... | .................... | ..................... | .................... | .................... | .................... |
| G | 3665 | hCoV-19/Australia/VIC3993/2020\|EPI_ISL_519383\|2020 | .................... | .................... | ..................... | .................... | .................... | .................... |
| G | 3666 | hCoV-19/Australia/VIC3995/2020\|EPI_ISL_519384\|2020 | .................... | .................... | ..................... | .................... | .................... | .................... |
| G | 3667 | hCoV-19/Australia/VIC3996/2020\|EPI_ISL_519385\|2020 | .................... | .................... | ..................... | .................... | .................... | .................... |
| G | 3668 | hCoV-19/Australia/VIC3997/2020\|EPI_ISL_519386\|2020 | .................... | .................... | ..................... | .................... | .................... | .................... |
| G | 3672 | hCoV-19/Australia/VIC4006/2020\|EPI_ISL_519392\|2020 | .................... | .................... | ..................... | .................... | .................... | .................... |
| G | 3673 | hCoV-19/Australia/VIC4007/2020\|EPI_ISL_519393\|2020 | .................... | .................... | ..................... | .................... | .................... | .................... |
| G | 3674 | hCoV-19/Australia/VIC4008/2020\|EPI_ISL_519394\|2020 | .................... | .................... | ..................... | .................... | .................... | .................... |
| G | 3675 | hCoV-19/Australia/VIC4009/2020\|EPI_ISL_519395\|2020 | .................... | .................... | ..................... | .................... | .................... | .................... |
| G | 3676 | hCoV-19/Australia/VIC4010/2020\|EPI_ISL_519396\|2020 | .................... | .................... | ..................... | .................... | .................... | .................... |
| G | 3677 | hCoV-19/Australia/VIC4011/2020\|EPI_ISL_519397\|2020 | .................... | .................... | ..................... | .................... | .................... | .................... |
| G | 3678 | hCoV-19/Australia/VIC4012/2020\|EPI_ISL_519398\|2020 | .................... | .................... | ..................... | .................... | .................... | .................... |
| G | 3679 | hCoV-19/Australia/VIC4013/2020\|EPI_ISL_519399\|2020 | .................... | .................... | ..................... | .................... | .................... | .................... |
| G | 3680 | hCoV-19/Australia/VIC4131/2020\|EPI_ISL_519503\|2020 | .................... | .................... | ..................... | .................... | .................... | .................... |
| G | 3681 | hCoV-19/Australia/VIC4132/2020\|EPI_ISL_519504\|2020 | .................... | .................... | ..................... | .................... | .................... | .................... |
| G | 3682 | hCoV-19/Australia/VIC4133/2020\|EPI_ISL_519505\|2020 | .................... | .................... | ..................... | .................... | .................... | .................... |
| G | 3683 | hCoV-19/Australia/VIC4134/2020\|EPI_ISL_519506\|2020 | .................... | .................... | ..................... | .................... | .................... | .................... |
| G | 3684 | hCoV-19/Australia/VIC4135/2020\|EPI_ISL_519507\|2020 | .................... | .................... | ..................... | .................... | .................... | .................... |
| G | 3685 | hCoV-19/Australia/VIC4136/2020\|EPI_ISL_519508\|2020 | .................... | .................... | ..................... | .................... | .................... | .................... |
| G | 3686 | hCoV-19/Australia/VIC4169/2020\|EPI_ISL_519518\|2020 | .................... | .................... | ..................... | .................... | .................... | .................... |
| G | 3687 | hCoV-19/Australia/VIC4179/2020\|EPI_ISL_519520\|2020 | .................... | .................... | ..................... | .................... | .................... | .................... |
| G | 3688 | hCoV-19/Australia/VIC4194/2020\|EPI_ISL_519522\|2020 | .................... | .................... | ..................... | .................... | .................... | .................... |
| G | 3689 | hCoV-19/Australia/VIC4195/2020\|EPI_ISL_519523\|2020 | .................... | .................... | ..................... | .................... | .................... | .................... |
| G | 3690 | hCoV-19/Australia/VIC4196/2020\|EPI_ISL_519524\|2020 | .................... | .................... | ..................... | .................... | .................... | .................... |
| G | 3691 | hCoV-19/Australia/VIC4197/2020\|EPI_ISL_519525\|2020 | .................... | .................... | ..................... | .................... | .................... | .................... |
| G | 3692 | hCoV-19/Australia/VIC4198/2020\|EPI_ISL_519526\|2020 | .................... | .................... | ..................... | .................... | .................... | .................... |
| G | 3693 | hCoV-19/Australia/VIC4199/2020\|EPI_ISL_519527\|2020 | .................... | .................... | ..................... | .................... | .................... | .................... |
| G | 3694 | hCoV-19/Australia/VIC4200/2020\|EPI_ISL_519528\|2020 | .................... | .................... | ..................... | .................... | .................... | .................... |
| G | 3695 | hCoV-19/Australia/VIC4201/2020\|EPI_ISL_519529\|2020 | .................... | .................... | ..................... | .................... | .................... | .................... |
| G | 3696 | hCoV-19/Australia/VIC4202/2020\|EPI_ISL_519530\|2020 | .................... | .................... | ..................... | .................... | .................... | .................... |
| G | 3697 | hCoV-19/Australia/VIC4203/2020\|EPI_ISL_519531\|2020 | .................... | .................... | ..................... | .................... | .................... | .................... |
| G | 3698 | hCoV-19/Australia/VIC4204/2020\|EPI_ISL_519532\|2020 | .................... | .................... | ..................... | .................... | .................... | .................... |
| G | 3699 | hCoV-19/Australia/VIC4205/2020\|EPI_ISL_519533\|2020 | .................... | .................... | ..................... | .................... | .................... | .................... |
| G | 3700 | hCoV-19/Australia/VIC4206/2020\|EPI_ISL_519534\|2020 | .................... | .................... | ..................... | .................... | .................... | .................... |
| G | 3701 | hCoV-19/Australia/VIC4207/2020\|EPI_ISL_519535\|2020 | .................... | .................... | ..................... | .................... | .................... | .................... |
| G | 3702 | hCoV-19/Australia/VIC4208/2020\|EPI_ISL_519536\|2020 | .................... | .................... | ..................... | .................... | .................... | .................... |
| G | 3703 | hCoV-19/Australia/VIC4209/2020\|EPI_ISL_519537\|2020 | .................... | .................... | ..................... | .................... | .................... | .................... |

<table>
<tr><td>G</td><td>3704</td><td>hCoV-19/Australia/VIC4210/2020|EPI_ISL_519538|2020</td><td>····················</td><td>····················</td><td>····················</td><td>····················</td><td>····················</td><td>····················</td><td></td></tr>
<tr><td>G</td><td>3705</td><td>hCoV-19/Australia/VIC4211/2020|EPI_ISL_519539|2020</td><td>····················</td><td>····················</td><td>····················</td><td>····················</td><td>····················</td><td>····················</td><td></td></tr>
<tr><td>G</td><td>3706</td><td>hCoV-19/Australia/VIC4212/2020|EPI_ISL_519540|2020</td><td>····················</td><td>····················</td><td>····················</td><td>····················</td><td>····················</td><td>····················</td><td></td></tr>
<tr><td>G</td><td>3707</td><td>hCoV-19/Australia/VIC4213/2020|EPI_ISL_519541|2020</td><td>····················</td><td>····················</td><td>····················</td><td>····················</td><td>····················</td><td>····················</td><td></td></tr>
<tr><td>G</td><td>3708</td><td>hCoV-19/Australia/VIC4214/2020|EPI_ISL_519542|2020</td><td>····················</td><td>····················</td><td>····················</td><td>····················</td><td>····················</td><td>····················</td><td></td></tr>
<tr><td>G</td><td>3709</td><td>hCoV-19/Australia/VIC4215/2020|EPI_ISL_519543|2020</td><td>····················</td><td>····················</td><td>····················</td><td>····················</td><td>····················</td><td>····················</td><td></td></tr>
<tr><td>G</td><td>3710</td><td>hCoV-19/Australia/VIC4216/2020|EPI_ISL_519544|2020</td><td>····················</td><td>····················</td><td>····················</td><td>····················</td><td>····················</td><td>····················</td><td></td></tr>
<tr><td>G</td><td>3711</td><td>hCoV-19/Australia/VIC4217/2020|EPI_ISL_519545|2020</td><td>····················</td><td>····················</td><td>····················</td><td>····················</td><td>····················</td><td>····················</td><td></td></tr>
<tr><td>G</td><td>3712</td><td>hCoV-19/Australia/VIC3594/2020|EPI_ISL_520340|2020</td><td>····················</td><td>····················</td><td>····················</td><td>····················</td><td>····················</td><td>····················</td><td></td></tr>
<tr><td>G</td><td>3713</td><td>hCoV-19/Australia/VIC3595/2020|EPI_ISL_520341|2020</td><td>····················</td><td>····················</td><td>····················</td><td>····················</td><td>····················</td><td>····················</td><td></td></tr>
<tr><td>G</td><td>3715</td><td>hCoV-19/Australia/VIC3598/2020|EPI_ISL_520344|2020</td><td>····················</td><td>····················</td><td>····················</td><td>····················</td><td>····················</td><td>····················</td><td></td></tr>
<tr><td>G</td><td>3718</td><td>hCoV-19/Australia/VIC3602/2020|EPI_ISL_520347|2020</td><td>····················</td><td>····················</td><td>····················</td><td>····················</td><td>····················</td><td>····················</td><td></td></tr>
<tr><td>G</td><td>3719</td><td>hCoV-19/Australia/VIC3604/2020|EPI_ISL_520349|2020</td><td>····················</td><td>····················</td><td>····················</td><td>····················</td><td>····················</td><td>····················</td><td></td></tr>
<tr><td>G</td><td>3721</td><td>hCoV-19/Australia/VIC3607/2020|EPI_ISL_520351|2020</td><td>····················</td><td>····················</td><td>····················</td><td>····················</td><td>····················</td><td>····················</td><td></td></tr>
<tr><td>G</td><td>3724</td><td>hCoV-19/Australia/VIC3610/2020|EPI_ISL_520354|2020</td><td>····················</td><td>····················</td><td>····················</td><td>····················</td><td>····················</td><td>····················</td><td></td></tr>
<tr><td>G</td><td>3726</td><td>hCoV-19/Australia/VIC3614/2020|EPI_ISL_520357|2020</td><td>····················</td><td>····················</td><td>····················</td><td>····················</td><td>····················</td><td>····················</td><td></td></tr>
<tr><td>G</td><td>3727</td><td>hCoV-19/Australia/VIC3615/2020|EPI_ISL_520358|2020</td><td>····················</td><td>····················</td><td>····················</td><td>····················</td><td>····················</td><td>····················</td><td></td></tr>
<tr><td>G</td><td>3728</td><td>hCoV-19/Australia/VIC3618/2020|EPI_ISL_520360|2020</td><td>····················</td><td>····················</td><td>····················</td><td>····················</td><td>····················</td><td>····················</td><td></td></tr>
<tr><td>G</td><td>3729</td><td>hCoV-19/Australia/VIC3619/2020|EPI_ISL_520361|2020</td><td>····················</td><td>····················</td><td>····················</td><td>····················</td><td>····················</td><td>····················</td><td></td></tr>
<tr><td>G</td><td>3730</td><td>hCoV-19/Australia/VIC3620/2020|EPI_ISL_520362|2020</td><td>····················</td><td>····················</td><td>····················</td><td>····················</td><td>····················</td><td>····················</td><td></td></tr>
<tr><td>G</td><td>3731</td><td>hCoV-19/Australia/VIC3621/2020|EPI_ISL_520363|2020</td><td>····················</td><td>····················</td><td>····················</td><td>····················</td><td>····················</td><td>····················</td><td></td></tr>
<tr><td>G</td><td>3732</td><td>hCoV-19/Australia/VIC3622/2020|EPI_ISL_520364|2020</td><td>····················</td><td>····················</td><td>····················</td><td>····················</td><td>····················</td><td>····················</td><td></td></tr>
<tr><td>G</td><td>3733</td><td>hCoV-19/Australia/VIC3623/2020|EPI_ISL_520365|2020</td><td>····················</td><td>····················</td><td>····················</td><td>····················</td><td>····················</td><td>····················</td><td></td></tr>
<tr><td>G</td><td>3734</td><td>hCoV-19/Australia/VIC3624/2020|EPI_ISL_520366|2020</td><td>····················</td><td>····················</td><td>····················</td><td>····················</td><td>····················</td><td>····················</td><td></td></tr>
<tr><td>G</td><td>3735</td><td>hCoV-19/Australia/VIC3626/2020|EPI_ISL_520367|2020</td><td>····················</td><td>····················</td><td>····················</td><td>····················</td><td>····················</td><td>····················</td><td></td></tr>
<tr><td>G</td><td>3736</td><td>hCoV-19/Australia/VIC3627/2020|EPI_ISL_520368|2020</td><td>····················</td><td>····················</td><td>····················</td><td>····················</td><td>NNN·················</td><td>····················</td><td>1</td></tr>
<tr><td>G</td><td>3737</td><td>hCoV-19/Australia/VIC3628/2020|EPI_ISL_520369|2020</td><td>····················</td><td>····················</td><td>····················</td><td>····················</td><td>····················</td><td>····················</td><td></td></tr>
<tr><td>G</td><td>3739</td><td>hCoV-19/Australia/VIC3630/2020|EPI_ISL_520371|2020</td><td>····················</td><td>····················</td><td>····················</td><td>····················</td><td>····················</td><td>····················</td><td></td></tr>
<tr><td>G</td><td>3742</td><td>hCoV-19/Australia/VIC3634/2020|EPI_ISL_520374|2020</td><td>····················</td><td>····················</td><td>····················</td><td>····················</td><td>····················</td><td>····················</td><td></td></tr>
<tr><td>G</td><td>3743</td><td>hCoV-19/Australia/VIC3635/2020|EPI_ISL_520375|2020</td><td>····················</td><td>····················</td><td>····················</td><td>····················</td><td>····················</td><td>····················</td><td></td></tr>
<tr><td>G</td><td>3745</td><td>hCoV-19/Australia/VIC3638/2020|EPI_ISL_520378|2020</td><td>····················</td><td>····················</td><td>····················</td><td>····················</td><td>····················</td><td>····················</td><td></td></tr>
<tr><td>G</td><td>3746</td><td>hCoV-19/Australia/VIC3639/2020|EPI_ISL_520379|2020</td><td>····················</td><td>····················</td><td>····················</td><td>····················</td><td>····················</td><td>····················</td><td></td></tr>
<tr><td>G</td><td>3747</td><td>hCoV-19/Australia/VIC3641/2020|EPI_ISL_520381|2020</td><td>····················</td><td>····················</td><td>····················</td><td>····················</td><td>····················</td><td>····················</td><td></td></tr>
<tr><td>G</td><td>3749</td><td>hCoV-19/Australia/VIC3645/2020|EPI_ISL_520385|2020</td><td>····················</td><td>····················</td><td>····················</td><td>····················</td><td>NNN·················</td><td>····················</td><td>1</td></tr>
<tr><td>G</td><td>3750</td><td>hCoV-19/Australia/VIC3647/2020|EPI_ISL_520386|2020</td><td>····················</td><td>····················</td><td>····················</td><td>····················</td><td>····················</td><td>····················</td><td></td></tr>
<tr><td>G</td><td>3751</td><td>hCoV-19/Australia/VIC3648/2020|EPI_ISL_520387|2020</td><td>····················</td><td>····················</td><td>····················</td><td>····················</td><td>····················</td><td>····················</td><td></td></tr>
<tr><td>G</td><td>3752</td><td>hCoV-19/Australia/VIC3649/2020|EPI_ISL_520388|2020</td><td>····················</td><td>····················</td><td>····················</td><td>····················</td><td>····················</td><td>····················</td><td></td></tr>
<tr><td>G</td><td>3753</td><td>hCoV-19/Australia/VIC3650/2020|EPI_ISL_520389|2020</td><td>····················</td><td>····················</td><td>····················</td><td>····················</td><td>····················</td><td>····················</td><td></td></tr>
<tr><td>G</td><td>3754</td><td>hCoV-19/Australia/VIC3651/2020|EPI_ISL_520390|2020</td><td>····················</td><td>····················</td><td>····················</td><td>····················</td><td>····················</td><td>····················</td><td></td></tr>
<tr><td>G</td><td>3755</td><td>hCoV-19/Australia/VIC3790/2020|EPI_ISL_520507|2020</td><td>····················</td><td>····················</td><td>····················</td><td>····················</td><td>····················</td><td>····················</td><td></td></tr>
<tr><td>G</td><td>3756</td><td>hCoV-19/Australia/VIC3791/2020|EPI_ISL_520508|2020</td><td>····················</td><td>····················</td><td>····················</td><td>····················</td><td>····················</td><td>····················</td><td></td></tr>
<tr><td>G</td><td>3757</td><td>hCoV-19/Australia/VIC3792/2020|EPI_ISL_520509|2020</td><td>····················</td><td>····················</td><td>····················</td><td>····················</td><td>····················</td><td>····················</td><td></td></tr>
<tr><td>G</td><td>3758</td><td>hCoV-19/Australia/VIC3793/2020|EPI_ISL_520510|2020</td><td>····················</td><td>····················</td><td>····················</td><td>····················</td><td>····················</td><td>····················</td><td></td></tr>
<tr><td>G</td><td>3759</td><td>hCoV-19/Australia/VIC3794/2020|EPI_ISL_520511|2020</td><td>····················</td><td>····················</td><td>····················</td><td>····················</td><td>····················</td><td>····················</td><td></td></tr>
<tr><td>G</td><td>3760</td><td>hCoV-19/Australia/VIC3795/2020|EPI_ISL_520512|2020</td><td>····················</td><td>····················</td><td>····················</td><td>····················</td><td>····················</td><td>····················</td><td></td></tr>
<tr><td>G</td><td>3761</td><td>hCoV-19/Australia/VIC3796/2020|EPI_ISL_520513|2020</td><td>····················</td><td>····················</td><td>····················</td><td>····················</td><td>····················</td><td>····················</td><td></td></tr>
<tr><td>G</td><td>3762</td><td>hCoV-19/Australia/VIC3799/2020|EPI_ISL_520516|2020</td><td>····················</td><td>····················</td><td>····················</td><td>····················</td><td>····················</td><td>····················</td><td></td></tr>
<tr><td>G</td><td>3763</td><td>hCoV-19/Australia/VIC3800/2020|EPI_ISL_520517|2020</td><td>····················</td><td>····················</td><td>····················</td><td>····················</td><td>····················</td><td>····················</td><td></td></tr>
<tr><td>G</td><td>3764</td><td>hCoV-19/Australia/VIC3801/2020|EPI_ISL_520518|2020</td><td>····················</td><td>····················</td><td>····················</td><td>····················</td><td>····················</td><td>····················</td><td></td></tr>
<tr><td>G</td><td>3765</td><td>hCoV-19/Australia/VIC3802/2020|EPI_ISL_520519|2020</td><td>····················</td><td>····················</td><td>····················</td><td>····················</td><td>····················</td><td>····················</td><td></td></tr>
<tr><td>G</td><td>3766</td><td>hCoV-19/Australia/VIC3803/2020|EPI_ISL_520520|2020</td><td>····················</td><td>····················</td><td>····················</td><td>····················</td><td>····················</td><td>····················</td><td></td></tr>
<tr><td>G</td><td>3767</td><td>hCoV-19/Australia/VIC3804/2020|EPI_ISL_520521|2020</td><td>····················</td><td>····················</td><td>····················</td><td>····················</td><td>····················</td><td>····················</td><td></td></tr>
<tr><td>G</td><td>3768</td><td>hCoV-19/Australia/VIC3809/2020|EPI_ISL_520526|2020</td><td>····················</td><td>····················</td><td>····················</td><td>····················</td><td>····················</td><td>····················</td><td></td></tr>
<tr><td>G</td><td>3769</td><td>hCoV-19/Australia/VIC2839/2020|EPI_ISL_521670|2020</td><td>····················</td><td>····················</td><td>····················</td><td>····················</td><td>····················</td><td>····················</td><td></td></tr>
<tr><td>G</td><td>3770</td><td>hCoV-19/Australia/VIC2840/2020|EPI_ISL_521671|2020</td><td>····················</td><td>····················</td><td>····················</td><td>····················</td><td>····················</td><td>····················</td><td></td></tr>
<tr><td>G</td><td>3771</td><td>hCoV-19/Australia/VIC2841/2020|EPI_ISL_521672|2020</td><td>····················</td><td>····················</td><td>····················</td><td>····················</td><td>····················</td><td>····················</td><td></td></tr>
<tr><td>G</td><td>3772</td><td>hCoV-19/Australia/VIC2842/2020|EPI_ISL_521673|2020</td><td>····················</td><td>····················</td><td>····················</td><td>····················</td><td>····················</td><td>····················</td><td></td></tr>
<tr><td>G</td><td>3773</td><td>hCoV-19/Australia/VIC2844/2020|EPI_ISL_521675|2020</td><td>····················</td><td>····················</td><td>····················</td><td>····················</td><td>····················</td><td>····················</td><td></td></tr>
<tr><td>G</td><td>3774</td><td>hCoV-19/Australia/VIC2845/2020|EPI_ISL_521676|2020</td><td>····················</td><td>····················</td><td>····················</td><td>····················</td><td>····················</td><td>····················</td><td></td></tr>
<tr><td>G</td><td>3775</td><td>hCoV-19/Australia/VIC2846/2020|EPI_ISL_521677|2020</td><td>····················</td><td>····················</td><td>····················</td><td>····················</td><td>····················</td><td>····················</td><td></td></tr>
<tr><td>G</td><td>3776</td><td>hCoV-19/Australia/VIC2847/2020|EPI_ISL_521678|2020</td><td>····················</td><td>····················</td><td>····················</td><td>····················</td><td>····················</td><td>····················</td><td></td></tr>
<tr><td>G</td><td>3777</td><td>hCoV-19/Australia/VIC2848/2020|EPI_ISL_521679|2020</td><td>····················</td><td>····················</td><td>····················</td><td>····················</td><td>····················</td><td>····················</td><td></td></tr>
<tr><td>G</td><td>3778</td><td>hCoV-19/Australia/VIC2849/2020|EPI_ISL_521680|2020</td><td>····················</td><td>····················</td><td>····················</td><td>····················</td><td>····················</td><td>····················</td><td></td></tr>
<tr><td>G</td><td>3779</td><td>hCoV-19/Australia/VIC2850/2020|EPI_ISL_521681|2020</td><td>····················</td><td>····················</td><td>····················</td><td>····················</td><td>····················</td><td>····················</td><td></td></tr>
<tr><td>G</td><td>3780</td><td>hCoV-19/Australia/VIC2852/2020|EPI_ISL_521683|2020</td><td>····················</td><td>····················</td><td>····················</td><td>····················</td><td>····················</td><td>····················</td><td></td></tr>
<tr><td>G</td><td>3781</td><td>hCoV-19/Australia/VIC2853/2020|EPI_ISL_521684|2020</td><td>····················</td><td>····················</td><td>····················</td><td>····················</td><td>····················</td><td>····················</td><td></td></tr>
<tr><td>G</td><td>3782</td><td>hCoV-19/Australia/VIC2854/2020|EPI_ISL_521685|2020</td><td>····················</td><td>····················</td><td>····················</td><td>····················</td><td>····················</td><td>····················</td><td></td></tr>
<tr><td>G</td><td>3783</td><td>hCoV-19/Australia/VIC2855/2020|EPI_ISL_521686|2020</td><td>····················</td><td>····················</td><td>····················</td><td>····················</td><td>····················</td><td>····················</td><td></td></tr>
<tr><td>G</td><td>3784</td><td>hCoV-19/Australia/VIC2856/2020|EPI_ISL_521687|2020</td><td>····················</td><td>····················</td><td>····················</td><td>····················</td><td>····················</td><td>····················</td><td></td></tr>
<tr><td>G</td><td>3785</td><td>hCoV-19/Australia/VIC2857/2020|EPI_ISL_521688|2020</td><td>····················</td><td>····················</td><td>····················</td><td>····················</td><td>····················</td><td>····················</td><td></td></tr>
<tr><td>G</td><td>3786</td><td>hCoV-19/Australia/VIC2858/2020|EPI_ISL_521689|2020</td><td>····················</td><td>····················</td><td>····················</td><td>····················</td><td>····················</td><td>····················</td><td></td></tr>
<tr><td>G</td><td>3787</td><td>hCoV-19/Australia/VIC2859/2020|EPI_ISL_521690|2020</td><td>····················</td><td>····················</td><td>····················</td><td>····················</td><td>····················</td><td>····················</td><td></td></tr>
<tr><td>G</td><td>3788</td><td>hCoV-19/Australia/VIC2860/2020|EPI_ISL_521691|2020</td><td>····················</td><td>····················</td><td>····················</td><td>····················</td><td>····················</td><td>····················</td><td></td></tr>
<tr><td>G</td><td>3789</td><td>hCoV-19/Australia/VIC2861/2020|EPI_ISL_521692|2020</td><td>····················</td><td>····················</td><td>····················</td><td>····················</td><td>····················</td><td>····················</td><td></td></tr>
<tr><td>G</td><td>3790</td><td>hCoV-19/Australia/VIC2862/2020|EPI_ISL_521693|2020</td><td>····················</td><td>····················</td><td>····················</td><td>····················</td><td>····················</td><td>····················</td><td></td></tr>
<tr><td>G</td><td>3791</td><td>hCoV-19/Australia/VIC2863/2020|EPI_ISL_521694|2020</td><td>····················</td><td>····················</td><td>····················</td><td>····················</td><td>····················</td><td>····················</td><td></td></tr>
<tr><td>G</td><td>3792</td><td>hCoV-19/Australia/VIC2864/2020|EPI_ISL_521695|2020</td><td>····················</td><td>····················</td><td>····················</td><td>····················</td><td>····················</td><td>····················</td><td></td></tr>
<tr><td>G</td><td>3793</td><td>hCoV-19/Australia/VIC2865/2020|EPI_ISL_521696|2020</td><td>····················</td><td>····················</td><td>····················</td><td>····················</td><td>····················</td><td>····················</td><td></td></tr>
<tr><td>G</td><td>3794</td><td>hCoV-19/Australia/VIC2866/2020|EPI_ISL_521697|2020</td><td>····················</td><td>····················</td><td>····················</td><td>····················</td><td>····················</td><td>····················</td><td></td></tr>
<tr><td>G</td><td>3795</td><td>hCoV-19/Australia/VIC2867/2020|EPI_ISL_521698|2020</td><td>····················</td><td>····················</td><td>····················</td><td>····················</td><td>····················</td><td>····················</td><td></td></tr>
</table>

| | | | | | | | | |
|---|---|---|---|---|---|---|---|---|
| G | 3796 hCoV-19/Australia/VIC2868/2020\|EPI_ISL_521699\|2020 | .................... | .................... | .................... | .................... | .................... | .................... | |
| G | 3797 hCoV-19/Australia/VIC2980/2020\|EPI_ISL_521810\|2020 | .................... | .................... | .................... | .................... | .................... | .................... | |
| G | 3798 hCoV-19/Australia/VIC2981/2020\|EPI_ISL_521811\|2020 | .................... | .................... | .................... | .................... | .................... | .................... | |
| G | 3799 hCoV-19/Australia/VIC2982/2020\|EPI_ISL_521812\|2020 | .................... | .................... | .................... | .................... | .................... | .................... | |
| G | 3800 hCoV-19/Australia/VIC2983/2020\|EPI_ISL_521813\|2020 | .................... | .................... | .................... | .................... | .................... | .................... | |
| G | 3801 hCoV-19/Australia/VIC2984/2020\|EPI_ISL_521814\|2020 | .................... | .................... | .................... | .................... | .................... | .................... | |
| G | 3802 hCoV-19/Australia/VIC2985/2020\|EPI_ISL_521815\|2020 | .................... | .................... | .................... | .................... | .................... | .................... | |
| G | 3803 hCoV-19/Australia/VIC2986/2020\|EPI_ISL_521816\|2020 | .................... | .................... | .................... | .................... | .................... | .................... | |
| G | 3804 hCoV-19/Australia/VIC2987/2020\|EPI_ISL_521817\|2020 | .................... | .................... | .................... | .................... | .................... | .................... | |
| G | 3805 hCoV-19/Australia/VIC2988/2020\|EPI_ISL_521818\|2020 | .................... | .................... | .................... | .................... | .................... | .................... | |
| G | 3806 hCoV-19/Australia/VIC2989/2020\|EPI_ISL_521819\|2020 | .................... | .................... | .................... | .................... | .................... | .................... | |
| G | 3807 hCoV-19/Australia/VIC2990/2020\|EPI_ISL_521820\|2020 | .................... | .................... | .................... | .................... | .................... | .................... | |
| G | 3808 hCoV-19/Australia/VIC2991/2020\|EPI_ISL_521821\|2020 | .................... | .................... | .................... | .................... | .................... | .................... | |
| G | 3809 hCoV-19/Australia/VIC2992/2020\|EPI_ISL_521822\|2020 | .................... | .................... | .................... | .................... | .................... | .................... | |
| G | 3810 hCoV-19/Australia/VIC2993/2020\|EPI_ISL_521823\|2020 | .................... | .................... | .................... | .................... | .................... | .................... | |
| G | 3811 hCoV-19/Australia/VIC2994/2020\|EPI_ISL_521824\|2020 | .................... | .................... | .................... | .................... | .................... | .................... | |
| G | 3812 hCoV-19/Australia/VIC2996/2020\|EPI_ISL_521826\|2020 | .................... | .................... | .................... | .................... | .................... | .................... | |
| G | 3813 hCoV-19/Australia/VIC2997/2020\|EPI_ISL_521827\|2020 | .................... | .................... | .................... | .................... | .................... | .................... | |
| G | 3814 hCoV-19/Australia/VIC2998/2020\|EPI_ISL_521828\|2020 | .................... | .................... | .................... | .................... | .................... | .................... | |
| G | 3815 hCoV-19/Australia/VIC2999/2020\|EPI_ISL_521829\|2020 | .................... | .................... | .................... | .................... | .................... | .................... | |
| G | 3816 hCoV-19/Australia/VIC3000/2020\|EPI_ISL_521830\|2020 | .................... | .................... | .................... | .................... | .................... | .................... | |
| G | 3817 hCoV-19/Australia/VIC3001/2020\|EPI_ISL_521831\|2020 | .................... | .................... | .................... | .................... | .................... | .................... | |
| G | 3818 hCoV-19/Australia/VIC3002/2020\|EPI_ISL_521832\|2020 | .................... | .................... | .................... | .................... | .................... | .................... | |
| G | 3819 hCoV-19/Australia/VIC3003/2020\|EPI_ISL_521833\|2020 | .................... | .................... | .................... | .................... | .................... | .................... | |
| G | 3820 hCoV-19/Australia/VIC3004/2020\|EPI_ISL_521834\|2020 | .................... | .................... | .................... | .................... | .................... | .................... | |
| G | 3821 hCoV-19/Australia/VIC3005/2020\|EPI_ISL_521835\|2020 | .................... | .................... | .................... | .................... | .................... | .................... | |
| G | 3822 hCoV-19/Australia/VIC3006/2020\|EPI_ISL_521836\|2020 | .................... | .................... | .................... | .................... | .................... | .................... | |
| G | 3823 hCoV-19/Australia/VIC3007/2020\|EPI_ISL_521837\|2020 | .................... | .................... | .................... | .................... | .................... | .................... | |
| G | 3824 hCoV-19/Australia/VIC3008/2020\|EPI_ISL_521838\|2020 | .................... | .................... | .................... | .................... | .................... | .................... | |
| G | 3825 hCoV-19/Australia/VIC3009/2020\|EPI_ISL_521839\|2020 | .................... | .................... | .................... | .................... | .................... | .................... | |
| G | 3826 hCoV-19/Australia/VIC3013/2020\|EPI_ISL_521842\|2020 | .................... | .................... | .................... | .................... | NNN................. | .................... | 1 |
| G | 3827 hCoV-19/Australia/VIC3014/2020\|EPI_ISL_521843\|2020 | .................... | .................... | .................... | .................... | .................... | .................... | |
| G | 3828 hCoV-19/Australia/VIC3015/2020\|EPI_ISL_521844\|2020 | .................... | .................... | .................... | .................... | .................... | .................... | |
| G | 3829 hCoV-19/Australia/VIC3016/2020\|EPI_ISL_521845\|2020 | .................... | .................... | .................... | .................... | .................... | .................... | |
| G | 3830 hCoV-19/Australia/VIC3018/2020\|EPI_ISL_521847\|2020 | .................... | .................... | .................... | .................... | .................... | .................... | |
| G | 3831 hCoV-19/Australia/VIC3020/2020\|EPI_ISL_521849\|2020 | .................... | .................... | .................... | .................... | .................... | .................... | |
| G | 3832 hCoV-19/Australia/VIC3021/2020\|EPI_ISL_521850\|2020 | .................... | .................... | .................... | .................... | .................... | .................... | |
| G | 3833 hCoV-19/Australia/VIC3022/2020\|EPI_ISL_521851\|2020 | .................... | .................... | .................... | .................... | .................... | .................... | |
| G | 3835 hCoV-19/Australia/VIC3028/2020\|EPI_ISL_521857\|2020 | .................... | .................... | .................... | .................... | .................... | .................... | |
| G | 3836 hCoV-19/Australia/VIC3029/2020\|EPI_ISL_521858\|2020 | .................... | .................... | .................... | .................... | .................... | .................... | |
| G | 3837 hCoV-19/USA/UT-08291/2020\|EPI_ISL_524090\|2020-05-1 | .................... | .................... | .................... | .................... | .................... | .................... | |
| G | 3838 hCoV-19/USA/UT-08296/2020\|EPI_ISL_524093\|2020-05-2 | .................... | .................... | .................... | .................... | .................... | .................... | |
| G | 3839 hCoV-19/USA/UT-08299/2020\|EPI_ISL_524096\|2020-05-2 | .................... | .................... | .................... | .................... | .................... | .................... | |
| G | 3840 hCoV-19/USA/UT-08301/2020\|EPI_ISL_524098\|2020-05-1 | .................... | .................... | .................... | .................... | .................... | .................... | |
| G | 3841 hCoV-19/Singapore/846/2020\|EPI_ISL_518010\|2020-07- | .................... | .................... | .................... | .................... | .................... | .................... | |
| G | 3842 hCoV-19/Singapore/847/2020\|EPI_ISL_518011\|2020-07- | .................... | .................... | .................... | .................... | .................... | .................... | |
| G | 3843 hCoV-19/Singapore/834_2/2020\|EPI_ISL_518013\|2020-0 | .................... | .................... | .................... | .................... | .................... | .................... | |
| G | 3844 hCoV-19/Singapore/849/2020\|EPI_ISL_518014\|2020-06- | .................... | .................... | .................... | .................... | .................... | .................... | |
| G | 3845 hCoV-19/Singapore/835_2/2020\|EPI_ISL_518015\|2020-0 | .................... | .................... | .................... | .................... | .................... | .................... | |
| G | 3846 hCoV-19/Singapore/836_2/2020\|EPI_ISL_518016\|2020-0 | .................... | .................... | .................... | .................... | .................... | .................... | |
| G | 3847 hCoV-19/Australia/VIC6050/2020\|EPI_ISL_518090\|2020 | .................... | .................... | .................... | .................... | .................... | .................... | |
| G | 3848 hCoV-19/Australia/VIC6051/2020\|EPI_ISL_518091\|2020 | .................... | .................... | .................... | .................... | .................... | .................... | |
| G | 3849 hCoV-19/Australia/VIC6052/2020\|EPI_ISL_518092\|2020 | .................... | .................... | .................... | .................... | .................... | .................... | |
| G | 3850 hCoV-19/Australia/VIC6054/2020\|EPI_ISL_518093\|2020 | .................... | .................... | .................... | .................... | .................... | .................... | |
| G | 3851 hCoV-19/Australia/VIC6055/2020\|EPI_ISL_518094\|2020 | .................... | .................... | .................... | .................... | .................... | .................... | |
| G | 3852 hCoV-19/Australia/VIC6058/2020\|EPI_ISL_518096\|2020 | .................... | .................... | .................... | .................... | .................... | .................... | |
| G | 3853 hCoV-19/Australia/VIC6059/2020\|EPI_ISL_518097\|2020 | .................... | .................... | .................... | .................... | .................... | .................... | |
| G | 3854 hCoV-19/Australia/VIC6070/2020\|EPI_ISL_518099\|2020 | .................... | .................... | .................... | .................... | .................... | .................... | |
| G | 3855 hCoV-19/Australia/VIC6105/2020\|EPI_ISL_518126\|2020 | .................... | .................... | .................... | .................... | .................... | .................... | |
| G | 3856 hCoV-19/Australia/VIC4974/2020\|EPI_ISL_519340\|2020 | .................... | .................... | .................... | .................... | .................... | .................... | |
| G | 3857 hCoV-19/Australia/VIC4975/2020\|EPI_ISL_519341\|2020 | .................... | .................... | .................... | .................... | .................... | .................... | |
| G | 3858 hCoV-19/Australia/VIC4976/2020\|EPI_ISL_519342\|2020 | .................... | .................... | .................... | .................... | .................... | .................... | |
| G | 3859 hCoV-19/Australia/VIC4977/2020\|EPI_ISL_519343\|2020 | .................... | .................... | .................... | .................... | .................... | .................... | |
| G | 3860 hCoV-19/Australia/VIC4978/2020\|EPI_ISL_519344\|2020 | .................... | .................... | .................... | .................... | .................... | .................... | |
| G | 3861 hCoV-19/Australia/VIC4979/2020\|EPI_ISL_519345\|2020 | .................... | .................... | .................... | .................... | .................... | .................... | |
| G | 3862 hCoV-19/Australia/VIC4980/2020\|EPI_ISL_519346\|2020 | .................... | .................... | .................... | .................... | .................... | .................... | |
| G | 3863 hCoV-19/Australia/VIC4981/2020\|EPI_ISL_519347\|2020 | .................... | .................... | .................... | .................... | .................... | .................... | |
| G | 3864 hCoV-19/Australia/VIC3953/2020\|EPI_ISL_519348\|2020 | .................... | .................... | .................... | .................... | .................... | .................... | |
| G | 3865 hCoV-19/Australia/VIC3955/2020\|EPI_ISL_519349\|2020 | .................... | .................... | .................... | .................... | .................... | .................... | |
| G | 3866 hCoV-19/Australia/VIC3958/2020\|EPI_ISL_519352\|2020 | .................... | .................... | .................... | .................... | .................... | .................... | |
| G | 3867 hCoV-19/Australia/VIC3959/2020\|EPI_ISL_519353\|2020 | .................... | .................... | .................... | .................... | .................... | .................... | |
| G | 3868 hCoV-19/Australia/VIC3961/2020\|EPI_ISL_519354\|2020 | .................... | .................... | .................... | .................... | .................... | .................... | |
| G | 3869 hCoV-19/Australia/VIC3962/2020\|EPI_ISL_519355\|2020 | .................... | .................... | .................... | .................... | .................... | .................... | |
| G | 3870 hCoV-19/Australia/VIC3965/2020\|EPI_ISL_519357\|2020 | .................... | .................... | .................... | .................... | .................... | .................... | |
| G | 3871 hCoV-19/Australia/VIC3966/2020\|EPI_ISL_519358\|2020 | .................... | .................... | .................... | .................... | .................... | .................... | |
| G | 3872 hCoV-19/Australia/VIC4076/2020\|EPI_ISL_519458\|2020 | .................... | .................... | .................... | .................... | .................... | .................... | |
| G | 3874 hCoV-19/Australia/VIC3577/2020\|EPI_ISL_520326\|2020 | .................... | .................... | .................... | .................... | .................... | .................... | |
| G | 3875 hCoV-19/Australia/VIC3578/2020\|EPI_ISL_520327\|2020 | .................... | .................... | .................... | .................... | .................... | .................... | |
| G | 3876 hCoV-19/Australia/VIC3580/2020\|EPI_ISL_520328\|2020 | .................... | .................... | .................... | .................... | .................... | .................... | |
| G | 3877 hCoV-19/Australia/VIC3581/2020\|EPI_ISL_520329\|2020 | .................... | .................... | .................... | .................... | .................... | .................... | |

| | | | | | | | | |
|---|---|---|---|---|---|---|---|---|
| G | 3878 hCoV-19/Australia/VIC3582/2020\|EPI_ISL_520330\|2020 | ···················· | ···················· | ···················· | ···················· | ···················· | ···················· | |
| G | 3880 hCoV-19/Australia/VIC3584/2020\|EPI_ISL_520332\|2020 | ···················· | ···················· | ···················· | ···················· | ···················· | ···················· | |
| G | 3881 hCoV-19/Australia/VIC3585/2020\|EPI_ISL_520333\|2020 | ···················· | ···················· | ···················· | ···················· | ···················· | ···················· | |
| G | 3882 hCoV-19/Australia/VIC3588/2020\|EPI_ISL_520334\|2020 | ···················· | ···················· | ···················· | ···················· | NNN················· | ···················· | 1 |
| G | 3883 hCoV-19/Australia/VIC3589/2020\|EPI_ISL_520335\|2020 | ···················· | ···················· | ···················· | ···················· | ···················· | ···················· | |
| G | 3884 hCoV-19/Australia/VIC3590/2020\|EPI_ISL_520336\|2020 | ···················· | ···················· | ···················· | ···················· | ···················· | ···················· | |
| G | 3885 hCoV-19/Australia/VIC3591/2020\|EPI_ISL_520337\|2020 | ···················· | ···················· | ···················· | ···················· | NNN················· | ···················· | 1 |
| G | 3887 hCoV-19/Australia/VIC3593/2020\|EPI_ISL_520339\|2020 | ···················· | ···················· | ···················· | ···················· | ···················· | ···················· | |
| G | 3889 hCoV-19/Australia/VIC2824/2020\|EPI_ISL_521655\|2020 | ···················· | ···················· | ···················· | ···················· | ···················· | ···················· | |
| G | 3890 hCoV-19/Australia/VIC2825/2020\|EPI_ISL_521656\|2020 | ···················· | ···················· | ···················· | ···················· | ···················· | ···················· | |
| G | 3891 hCoV-19/Australia/VIC2826/2020\|EPI_ISL_521657\|2020 | ···················· | ···················· | ···················· | ···················· | ···················· | ···················· | |
| G | 3892 hCoV-19/Australia/VIC2827/2020\|EPI_ISL_521658\|2020 | ···················· | ···················· | ···················· | ···················· | ···················· | ···················· | |
| G | 3893 hCoV-19/Australia/VIC2828/2020\|EPI_ISL_521659\|2020 | ···················· | ···················· | ···················· | ···················· | ···················· | ···················· | |
| G | 3894 hCoV-19/Australia/VIC2829/2020\|EPI_ISL_521660\|2020 | ···················· | ···················· | ···················· | ···················· | ···················· | ···················· | |
| G | 3895 hCoV-19/Australia/VIC2830/2020\|EPI_ISL_521661\|2020 | ···················· | ···················· | ···················· | ···················· | ···················· | ···················· | |
| G | 3896 hCoV-19/Australia/VIC2831/2020\|EPI_ISL_521662\|2020 | ···················· | ···················· | ···················· | ···················· | ···················· | ···················· | |
| G | 3897 hCoV-19/Australia/VIC2832/2020\|EPI_ISL_521663\|2020 | ···················· | ···················· | ···················· | ···················· | ···················· | ···················· | |
| G | 3898 hCoV-19/Australia/VIC2833/2020\|EPI_ISL_521664\|2020 | ···················· | ···················· | ···················· | ···················· | ···················· | ···················· | |
| G | 3899 hCoV-19/Australia/VIC2834/2020\|EPI_ISL_521665\|2020 | ···················· | ···················· | ···················· | ···················· | ···················· | ···················· | |
| G | 3900 hCoV-19/Australia/VIC2835/2020\|EPI_ISL_521666\|2020 | ···················· | ···················· | ···················· | ···················· | ···················· | ···················· | |
| G | 3901 hCoV-19/Australia/VIC2836/2020\|EPI_ISL_521667\|2020 | ···················· | ···················· | ···················· | ···················· | ···················· | ···················· | |
| G | 3902 hCoV-19/Australia/VIC2837/2020\|EPI_ISL_521668\|2020 | ···················· | ···················· | ···················· | ···················· | ···················· | ···················· | |
| G | 3903 hCoV-19/Australia/VIC2838/2020\|EPI_ISL_521669\|2020 | ···················· | ···················· | ···················· | ···················· | ···················· | ···················· | |
| G | 3904 hCoV-19/Australia/VIC2843/2020\|EPI_ISL_521674\|2020 | ···················· | ···················· | ···················· | ···················· | ···················· | ···················· | |
| G | 3905 hCoV-19/Australia/VIC2936/2020\|EPI_ISL_521767\|2020 | ···················· | ···················· | ···················· | ···················· | ···················· | ···················· | |
| G | 3906 hCoV-19/Australia/VIC2937/2020\|EPI_ISL_521768\|2020 | ···················· | ···················· | ···················· | ···················· | ···················· | ···················· | |
| G | 3907 hCoV-19/Australia/VIC2940/2020\|EPI_ISL_521770\|2020 | ···················· | ···················· | ···················· | ···················· | ···················· | ···················· | |
| G | 3908 hCoV-19/Australia/VIC2941/2020\|EPI_ISL_521771\|2020 | ···················· | ···················· | ···················· | ···················· | ···················· | ···················· | |
| G | 3909 hCoV-19/Australia/VIC2942/2020\|EPI_ISL_521772\|2020 | ···················· | ···················· | ···················· | ···················· | ···················· | ···················· | |
| G | 3910 hCoV-19/Australia/VIC2943/2020\|EPI_ISL_521773\|2020 | ···················· | ···················· | ···················· | ···················· | ···················· | ···················· | |
| G | 3911 hCoV-19/Australia/VIC2944/2020\|EPI_ISL_521774\|2020 | ···················· | ···················· | ···················· | ···················· | ···················· | ···················· | |
| G | 3913 hCoV-19/mink/Netherlands/NB-EMC-10-13/2020\|EPI_ISL | ···················· | ···················· | ···················· | ···················· | ···················· | ···················· | |
| G | 3914 hCoV-19/mink/Netherlands/NB-EMC-10-2/2020\|EPI_ISL_ | ···················· | ···················· | ···················· | ···················· | ···················· | ···················· | |
| G | 3915 hCoV-19/mink/Netherlands/NB-EMC-10-5/2020\|EPI_ISL_ | ···················· | ···················· | ···················· | ···················· | ···················· | ···················· | |
| G | 3916 hCoV-19/mink/Netherlands/NB-EMC-10-8/2020\|EPI_ISL_ | ···················· | ···················· | ···················· | ···················· | ···················· | ···················· | |
| G | 3917 hCoV-19/mink/Netherlands/NB-EMC-1-10/2020\|EPI_ISL_ | ···················· | ···················· | ···················· | ···················· | ···················· | ···················· | |
| G | 3918 hCoV-19/mink/Netherlands/NB-EMC-1-11/2020\|EPI_ISL_ | ···················· | ···················· | ···················· | ···················· | ···················· | ···················· | |
| G | 3919 hCoV-19/mink/Netherlands/NB-EMC-11-19/2020\|EPI_ISL | ···················· | ···················· | ···················· | ···················· | ···················· | ···················· | |
| G | 3920 hCoV-19/mink/Netherlands/NB-EMC-11-2/2020\|EPI_ISL_ | ···················· | ···················· | ···················· | ···················· | ···················· | ···················· | |
| G | 3921 hCoV-19/mink/Netherlands/NB-EMC-1-12/2020\|EPI_ISL_ | ···················· | ···················· | ···················· | ···················· | ···················· | ···················· | |
| G | 3922 hCoV-19/mink/Netherlands/NB-EMC-11-3/2020\|EPI_ISL_ | ···················· | ···················· | ···················· | ···················· | ···················· | ···················· | |
| G | 3923 hCoV-19/mink/Netherlands/NB-EMC-1-13/2020\|EPI_ISL_ | ···················· | ···················· | ···················· | ···················· | ···················· | ···················· | |
| G | 3924 hCoV-19/USA/VA-DCLS-1173/2020\|EPI_ISL_525200\|2020- | ···················· | ·A·················· | ···················· | ···················· | ···················· | ···················· | |
| G | 3925 hCoV-19/USA/VA-DCLS-1174/2020\|EPI_ISL_525201\|2020- | ···················· | ·A·················· | ···················· | ···················· | ···················· | ···················· | |
| G | 3926 hCoV-19/USA/VA-DCLS-1175/2020\|EPI_ISL_525202\|2020- | ···················· | ···················· | ···················· | ···················· | ···················· | ···················· | |
| G | 3927 hCoV-19/USA/VA-DCLS-1176/2020\|EPI_ISL_525203\|2020- | ···················· | ···················· | ···················· | ···················· | ···················· | ···················· | |
| G | 3928 hCoV-19/USA/VA-DCLS-1177/2020\|EPI_ISL_525204\|2020- | ···················· | ···················· | ···················· | ···················· | ···················· | ···················· | |
| G | 3929 hCoV-19/USA/VA-DCLS-1178/2020\|EPI_ISL_525205\|2020- | ···················· | ···················· | ···················· | ···················· | ···················· | ···················· | |
| G | 3930 hCoV-19/Peru/LAL-INS-064/2020\|EPI_ISL_525206\|2020- | ···················· | ···················· | ···················· | ···················· | ···················· | ···················· | |
| G | 3931 hCoV-19/USA/UT-UPHL-2009208/2020\|EPI_ISL_525208\|20 | ···················· | ···················· | ···················· | ···················· | ···················· | ···················· | |
| G | 3932 hCoV-19/USA/UT-UPHL-2009209/2020\|EPI_ISL_525209\|20 | ···················· | ···················· | ···················· | ···················· | ···················· | ···················· | |
| G | 3933 hCoV-19/USA/UT-UPHL-2009211/2020\|EPI_ISL_525211\|20 | ···················· | ···················· | ···················· | ···················· | ···················· | ···················· | |
| G | 3934 hCoV-19/USA/UT-UPHL-2009213/2020\|EPI_ISL_525213\|20 | ···················· | ···················· | ···················· | ···················· | ···················· | ···················· | |
| G | 3935 hCoV-19/USA/UT-UPHL-2009215/2020\|EPI_ISL_525215\|20 | ···················· | ···················· | ···················· | ···················· | ···················· | ···················· | |
| G | 3936 hCoV-19/USA/UT-UPHL-2009216/2020\|EPI_ISL_525216\|20 | ···················· | ···················· | ···················· | ···················· | ···················· | ···················· | |
| G | 3937 hCoV-19/USA/UT-UPHL-2009217/2020\|EPI_ISL_525217\|20 | ···················· | ···················· | ···················· | ···················· | ··C················· | ···················· | |
| G | 3938 hCoV-19/USA/UT-UPHL-2009219/2020\|EPI_ISL_525219\|20 | ···················· | ···················· | ···················· | ···················· | ···················· | ···················· | |
| G | 3939 hCoV-19/USA/UT-UPHL-2009221/2020\|EPI_ISL_525221\|20 | ···················· | ···················· | ···················· | ···················· | ···················· | ···················· | |
| G | 3940 hCoV-19/USA/UT-UPHL-2009223/2020\|EPI_ISL_525223\|20 | ···················· | ···················· | ···················· | ···················· | ···················· | ···················· | |
| G | 3941 hCoV-19/USA/UT-UPHL-2009225/2020\|EPI_ISL_525224\|20 | ···················· | ···················· | ···················· | ···················· | ···················· | ···················· | |
| G | 3942 hCoV-19/USA/UT-UPHL-2009227/2020\|EPI_ISL_525225\|20 | ···················· | ···················· | ···················· | ···················· | ···················· | ···················· | |
| G | 3943 hCoV-19/USA/UT-UPHL-2009231/2020\|EPI_ISL_525227\|20 | ···················· | ···················· | ···················· | ···················· | ·········A·········· | ···················· | |
| G | 3944 hCoV-19/USA/UT-UPHL-2009235/2020\|EPI_ISL_525228\|20 | ···················· | ···················· | ···················· | ···················· | ···················· | ···················· | |
| G | 3945 hCoV-19/USA/UT-UPHL-2009236/2020\|EPI_ISL_525229\|20 | ···················· | ···················· | ···················· | ···················· | ···················· | ···················· | |
| G | 3946 hCoV-19/USA/UT-UPHL-2009239/2020\|EPI_ISL_525231\|20 | ···················· | ···················· | ···················· | ···················· | ···················· | ···················· | |
| G | 3947 hCoV-19/USA/UT-UPHL-2009244/2020\|EPI_ISL_525233\|20 | ···················· | ···················· | ···················· | ···················· | ·········A·········· | ···················· | |
| G | 3948 hCoV-19/USA/UT-UPHL-2009248/2020\|EPI_ISL_525236\|20 | ···················· | ···················· | ···················· | ···················· | ···················· | ···················· | |
| G | 3949 hCoV-19/USA/UT-UPHL-2009249/2020\|EPI_ISL_525237\|20 | ···················· | ···················· | ···················· | ···················· | ···················· | ···················· | |
| G | 3950 hCoV-19/USA/UT-UPHL-2009252/2020\|EPI_ISL_525239\|20 | ···················· | ···················· | ···················· | ···················· | ···················· | ···················· | |
| G | 3951 hCoV-19/USA/UT-UPHL-2009255/2020\|EPI_ISL_525241\|20 | ···················· | ···················· | ···················· | ···················· | ···················· | ···················· | |
| G | 3952 hCoV-19/USA/UT-UPHL-2009256/2020\|EPI_ISL_525242\|20 | ···················· | ···················· | ···················· | ···················· | ···················· | ···················· | |
| G | 3953 hCoV-19/USA/UT-UPHL-2009260/2020\|EPI_ISL_525244\|20 | ···················· | ···················· | ···················· | ···················· | ···················· | ···················· | |
| G | 3954 hCoV-19/USA/UT-UPHL-2009262/2020\|EPI_ISL_525246\|20 | ···················· | ···················· | ···················· | ···················· | ···················· | ···················· | |
| G | 3955 hCoV-19/USA/UT-UPHL-2009264/2020\|EPI_ISL_525247\|20 | ···················· | ···················· | ···················· | ···················· | ···················· | ···················· | |
| G | 3956 hCoV-19/USA/UT-UPHL-2009265/2020\|EPI_ISL_525248\|20 | ···················· | ···················· | ···················· | ···················· | ···················· | ···················· | |
| G | 3957 hCoV-19/USA/UT-UPHL-2009267/2020\|EPI_ISL_525250\|20 | ···················· | ···················· | ···················· | ···················· | ···················· | ···················· | |
| G | 3958 hCoV-19/USA/UT-UPHL-2009268/2020\|EPI_ISL_525251\|20 | ···················· | ···················· | ···················· | ···················· | ···················· | ···················· | |
| G | 3959 hCoV-19/USA/UT-UPHL-2009269/2020\|EPI_ISL_525252\|20 | ···················· | ···················· | ···················· | ···················· | ···················· | ···················· | |
| G | 3960 hCoV-19/USA/UT-UPHL-2009274/2020\|EPI_ISL_525255\|20 | ···················· | ···················· | ···················· | ···················· | ···················· | ···················· | |
| G | 3961 hCoV-19/USA/UT-UPHL-2009275/2020\|EPI_ISL_525256\|20 | ···················· | ···················· | ···················· | ···················· | ···················· | ···················· | |

| | | | | | | | | |
|---|---|---|---|---|---|---|---|---|
| G | 3962 | hCoV-19/USA/UT-UPHL-2009277/2020\|EPI_ISL_525258\|20 | ···················· | ···················· | ····················· | ···················· | ···················· | ···················· |
| G | 3963 | hCoV-19/USA/UT-UPHL-2009278/2020\|EPI_ISL_525259\|20 | ···················· | ···················· | ····················· | ···················· | ···················· | ···················· |
| G | 3964 | hCoV-19/USA/UT-UPHL-2009281/2020\|EPI_ISL_525262\|20 | ···················· | ···················· | ····················· | ···················· | ···················· | ···················· |
| G | 3965 | hCoV-19/USA/UT-UPHL-2009282/2020\|EPI_ISL_525263\|20 | ···················· | ···················· | ····················· | ···················· | ···················· | ···················· |
| G | 3966 | hCoV-19/USA/UT-UPHL-2009286/2020\|EPI_ISL_525265\|20 | ···················· | ···················· | ····················· | ···················· | ··C················ | ···················· |
| G | 3967 | hCoV-19/USA/UT-UPHL-2009287/2020\|EPI_ISL_525266\|20 | ···················· | ···················· | ····················· | ···················· | ···················· | ···················· |
| G | 3968 | hCoV-19/USA/UT-UPHL-2009290/2020\|EPI_ISL_525268\|20 | ···················· | ···················· | ····················· | ···················· | ···················· | ···················· |
| G | 3969 | hCoV-19/USA/UT-UPHL-2009292/2020\|EPI_ISL_525270\|20 | ···················· | ···················· | ····················· | ···················· | ···················· | ···················· |
| G | 3970 | hCoV-19/USA/UT-UPHL-2009293/2020\|EPI_ISL_525271\|20 | ···················· | ···················· | ····················· | ···················· | ···················· | ···················· |
| G | 3971 | hCoV-19/USA/UT-UPHL-2008944/2020\|EPI_ISL_525273\|20 | ···················· | ···················· | ····················· | ···················· | ···················· | ···················· |
| G | 3972 | hCoV-19/USA/UT-UPHL-2008948/2020\|EPI_ISL_525275\|20 | ···················· | ···················· | ····················· | ···················· | ···················· | ···················· |
| G | 3973 | hCoV-19/USA/UT-UPHL-2008950/2020\|EPI_ISL_525277\|20 | ···················· | ···················· | ····················· | ···················· | ···················· | ···················· |
| G | 3974 | hCoV-19/USA/UT-UPHL-2008953/2020\|EPI_ISL_525280\|20 | ···················· | ···················· | ····················· | ···················· | ···················· | ···················· |
| G | 3975 | hCoV-19/USA/UT-UPHL-2008954/2020\|EPI_ISL_525281\|20 | ···················· | ···················· | ····················· | ···················· | ···················· | ···················· |
| G | 3976 | hCoV-19/USA/UT-UPHL-2008955/2020\|EPI_ISL_525282\|20 | ···················· | ···················· | ····················· | ···················· | ···················· | ···················· |
| G | 3977 | hCoV-19/USA/UT-UPHL-2008956/2020\|EPI_ISL_525283\|20 | ···················· | ···················· | ····················· | ···················· | ···················· | ···················· |
| G | 3978 | hCoV-19/USA/UT-UPHL-2008957/2020\|EPI_ISL_525284\|20 | ···················· | ···················· | ····················· | ···················· | ···················· | ···················· |
| G | 3979 | hCoV-19/USA/UT-UPHL-2008959/2020\|EPI_ISL_525286\|20 | ···················· | ···················· | ····················· | ···················· | ···················· | ···················· |
| G | 3980 | hCoV-19/USA/UT-UPHL-2008960/2020\|EPI_ISL_525287\|20 | ···················· | ···················· | ····················· | ···················· | ···················· | ···················· |
| G | 3981 | hCoV-19/USA/UT-UPHL-2008962/2020\|EPI_ISL_525288\|20 | ···················· | ···················· | ····················· | ···················· | ···················· | ···················· |
| G | 3982 | hCoV-19/USA/UT-UPHL-2008964/2020\|EPI_ISL_525290\|20 | ···················· | ···················· | ····················· | ···················· | ···················· | ···················· |
| G | 3983 | hCoV-19/USA/UT-UPHL-2008965/2020\|EPI_ISL_525291\|20 | ···················· | ···················· | ····················· | ···················· | ···················· | ···················· |
| G | 3984 | hCoV-19/USA/UT-UPHL-2008968/2020\|EPI_ISL_525292\|20 | ···················· | ···················· | ····················· | ···················· | ···················· | ···················· |
| G | 3985 | hCoV-19/USA/UT-UPHL-2008969/2020\|EPI_ISL_525293\|20 | ···················· | ···················· | ····················· | ···················· | ···················· | ···················· |
| G | 3986 | hCoV-19/USA/UT-UPHL-2008971/2020\|EPI_ISL_525295\|20 | ···················· | ···················· | ····················· | ···················· | ···················· | ···················· |
| G | 3987 | hCoV-19/USA/UT-UPHL-2008975/2020\|EPI_ISL_525297\|20 | ···················· | ···················· | ····················· | ···················· | ···················· | ···················· |
| G | 3988 | hCoV-19/USA/UT-UPHL-2008979/2020\|EPI_ISL_525299\|20 | ···················· | ···················· | ····················· | ···················· | ···················· | ···················· |
| G | 3989 | hCoV-19/Scotland/EDB6886/2020\|EPI_ISL_526500\|2020- | ···················· | ·A·················· | ····················· | ···················· | ···················· | ···················· |
| G | 3990 | hCoV-19/Scotland/EDB6887/2020\|EPI_ISL_526501\|2020- | ···················· | ·A·················· | ····················· | ···················· | ···················· | ···················· |
| G | 3991 | hCoV-19/Scotland/EDB6888/2020\|EPI_ISL_526502\|2020- | ···················· | ·A·················· | ····················· | ···················· | ···················· | ···················· |
| G | 3992 | hCoV-19/Scotland/EDB6893/2020\|EPI_ISL_526503\|2020- | ···················· | ·A·················· | ····················· | ···················· | ···················· | ···················· |
| G | 3993 | hCoV-19/Scotland/EDB6894/2020\|EPI_ISL_526504\|2020- | ···················· | ·A·················· | ····················· | ···················· | ···················· | ···················· |
| G | 3994 | hCoV-19/Scotland/EDB6895/2020\|EPI_ISL_526505\|2020- | ···················· | ·A·················· | ····················· | ···················· | ···················· | ···················· |
| G | 3995 | hCoV-19/Scotland/EDB6897/2020\|EPI_ISL_526506\|2020- | ···················· | ·A·················· | ····················· | ···················· | ···················· | ···················· |
| G | 3996 | hCoV-19/Scotland/EDB6898/2020\|EPI_ISL_526507\|2020- | ···················· | ·A·················· | ····················· | ···················· | ···················· | ···················· |
| G | 3997 | hCoV-19/Scotland/EDB6899/2020\|EPI_ISL_526508\|2020- | ···················· | ·A·················· | ····················· | ···················· | ···················· | ···················· |
| G | 3998 | hCoV-19/Scotland/EDB6903/2020\|EPI_ISL_526509\|2020- | ···················· | ·A·················· | ····················· | ···················· | ···················· | ···················· |
| G | 3999 | hCoV-19/Scotland/EDB6904/2020\|EPI_ISL_526510\|2020- | ···················· | ·A·················· | ····················· | ···················· | ···················· | ···················· |
| G | 4000 | hCoV-19/Scotland/EDB6905/2020\|EPI_ISL_526511\|2020- | ···················· | ·A·················· | ····················· | ···················· | ···················· | ···················· |
| G | 4001 | hCoV-19/Scotland/EDB6906/2020\|EPI_ISL_526512\|2020- | ···················· | ·A·················· | ····················· | ···················· | ···················· | ···················· |
| G | 4002 | hCoV-19/Scotland/EDB6907/2020\|EPI_ISL_526513\|2020- | ···················· | ·A·················· | ····················· | ···················· | ···················· | ···················· |
| G | 4003 | hCoV-19/Scotland/EDB6909/2020\|EPI_ISL_526514\|2020- | ···················· | ·A·················· | ····················· | ···················· | ···················· | ···················· |
| G | 4004 | hCoV-19/Scotland/EDB6911/2020\|EPI_ISL_526515\|2020- | ···················· | ·A·················· | ····················· | ···················· | ···················· | ···················· |
| G | 4007 | hCoV-19/Scotland/EDB7358/2020\|EPI_ISL_526532\|2020- | ···················· | ···················· | ····················· | ···················· | ···················· | ···················· |
| G | 4008 | hCoV-19/Scotland/EDB7359/2020\|EPI_ISL_526533\|2020- | ···················· | ···················· | ····················· | ···················· | ···················· | ···················· |
| G | 4009 | hCoV-19/Scotland/EDB7360/2020\|EPI_ISL_526534\|2020- | ···················· | ···················· | ····················· | ···················· | ···················· | ···················· |
| G | 4011 | hCoV-19/England/20164005604/2020\|EPI_ISL_526537\|20 | ···················· | ···················· | ····················· | ···················· | ···················· | ···················· |
| G | 4012 | hCoV-19/England/20164006104/2020\|EPI_ISL_526538\|20 | ···················· | ···················· | ····················· | ···················· | ···················· | ···················· |
| G | 4013 | hCoV-19/England/20164006804/2020\|EPI_ISL_526539\|20 | ···················· | ···················· | ····················· | ···················· | ···················· | ···················· |
| G | 4014 | hCoV-19/England/20192146704/2020\|EPI_ISL_526545\|20 | ···················· | ···················· | ····················· | ···················· | ···················· | ···················· |
| G | 4015 | hCoV-19/England/20198250304/2020\|EPI_ISL_526547\|20 | ···················· | ···················· | ····················· | ···················· | ···················· | ···················· |
| G | 4016 | hCoV-19/England/20204112904/2020\|EPI_ISL_526550\|20 | ···················· | ···················· | ····················· | ···················· | ···················· | ···················· |
| G | 4017 | hCoV-19/USA/OR-OHSU-1595/2020\|EPI_ISL_526551\|2020- | ···················· | ···················· | ····················· | ···················· | ···················· | ···················· |
| G | 4018 | hCoV-19/USA/FL-BPHL-0884/2020\|EPI_ISL_526553\|2020- | ···················· | ···················· | ····················· | ···················· | ···················· | ···················· |
| G | 4019 | hCoV-19/USA/FL-BPHL-1069/2020\|EPI_ISL_526555\|2020- | ···················· | ···················· | ····················· | ···················· | ···················· | ···················· |
| G | 4020 | hCoV-19/USA/FL-BPHL-1077/2020\|EPI_ISL_526563\|2020- | ···················· | ···················· | ····················· | ···················· | ···················· | ···················· |
| G | 4022 | hCoV-19/USA/FL-BPHL-1082/2020\|EPI_ISL_526568\|2020- | ···················· | ···················· | ····················· | ···················· | ···················· | ···················· |
| G | 4023 | hCoV-19/USA/FL-BPHL-1083/2020\|EPI_ISL_526569\|2020- | ···················· | ···················· | ····················· | ···················· | ···················· | ···················· |
| G | 4025 | hCoV-19/USA/FL-BPHL-1085/2020\|EPI_ISL_526571\|2020- | ···················· | ···················· | ····················· | ···················· | ···················· | ···················· |
| G | 4026 | hCoV-19/USA/FL-BPHL-1086/2020\|EPI_ISL_526572\|2020- | ···················· | ···················· | ····················· | ···················· | ···················· | ···················· |
| G | 4027 | hCoV-19/USA/FL-BPHL-1087/2020\|EPI_ISL_526573\|2020- | ···················· | ···················· | ····················· | ···················· | ···················· | ···················· |
| G | 4028 | hCoV-19/USA/FL-BPHL-1089/2020\|EPI_ISL_526575\|2020- | ···················· | ···················· | ····················· | ···················· | ···················· | ···············A···· |
| G | 4029 | hCoV-19/USA/FL-BPHL-1094/2020\|EPI_ISL_526580\|2020- | ···················· | ···················· | ····················· | ···················· | ···················· | ···················· |
| G | 4031 | hCoV-19/USA/FL-BPHL-1097/2020\|EPI_ISL_526583\|2020- | ···················· | ····A··············· | ····················· | ···················· | ···················· | ···················· |
| G | 4032 | hCoV-19/USA/FL-BPHL-1099/2020\|EPI_ISL_526585\|2020- | ···················· | ···················· | ····················· | ···················· | ···················· | ···················· |
| G | 4033 | hCoV-19/USA/FL-BPHL-1100/2020\|EPI_ISL_526586\|2020- | ···················· | ···················· | ····················· | ···················· | ···················· | ···················· |
| G | 4035 | hCoV-19/USA/FL-BPHL-1103/2020\|EPI_ISL_526589\|2020- | ···················· | ···················· | ····················· | ···················· | ···················· | ···················· |
| G | 4036 | hCoV-19/USA/FL-BPHL-1104/2020\|EPI_ISL_526590\|2020- | ···················· | ···················· | ····················· | ···················· | ···················· | ···················· |
| G | 4037 | hCoV-19/USA/FL-BPHL-1105/2020\|EPI_ISL_526591\|2020- | ···················· | ···················· | ····················· | ···················· | ···················· | ···················· |
| G | 4038 | hCoV-19/USA/FL-BPHL-1106/2020\|EPI_ISL_526592\|2020- | ···················· | ···················· | ····················· | ···················· | ···················· | ···················· |
| G | 4039 | hCoV-19/USA/FL-BPHL-1107/2020\|EPI_ISL_526593\|2020- | ···················· | ···················· | ····················· | ···················· | ···················· | ···················· |
| G | 4040 | hCoV-19/USA/FL-BPHL-1108/2020\|EPI_ISL_526594\|2020- | ···················· | ···················· | ····················· | ···················· | ···················· | ···················· |
| G | 4041 | hCoV-19/USA/FL-BPHL-1109/2020\|EPI_ISL_526595\|2020- | ···················· | ···················· | ····················· | ···················· | ···················· | ···················· |
| G | 4042 | hCoV-19/USA/FL-BPHL-1110/2020\|EPI_ISL_526596\|2020- | ···················· | ···················· | ····················· | ···················· | ···················· | ···················· |
| G | 4043 | hCoV-19/USA/FL-BPHL-1111/2020\|EPI_ISL_526597\|2020- | ···················· | ···················· | ····················· | ···················· | ···················· | ···················· |
| G | 4044 | hCoV-19/USA/FL-BPHL-1112/2020\|EPI_ISL_526598\|2020- | ···················· | ···················· | ····················· | ···················· | ···················· | ···················· |
| G | 4045 | hCoV-19/USA/FL-BPHL-1113/2020\|EPI_ISL_526599\|2020- | ···················· | ···················· | ····················· | ···················· | ···················· | ···················· |
| G | 4052 | hCoV-19/Ecuador/USFQ-193/2020\|EPI_ISL_527809\|2020- | ···················· | ···················· | ····················· | ···················· | ···················· | ···················· |
| G | 4053 | hCoV-19/Ecuador/USFQ-194/2020\|EPI_ISL_527810\|2020- | ···················· | ···················· | ····················· | ···················· | ···················· | ···················· |
| G | 4054 | hCoV-19/Ecuador/USFQ-195/2020\|EPI_ISL_527811\|2020- | ···················· | ···················· | ····················· | ···················· | ···················· | ···················· |

| | | | | | | | | |
|---|---|---|---|---|---|---|---|---|
| G | 4055 | hCoV-19/Ecuador/USFQ-196/2020\|EPI_ISL_527812\|2020- | ···················· | ···················· | ···················· | ···················· | ···················· | ···················· |
| G | 4056 | hCoV-19/Ecuador/USFQ-197/2020\|EPI_ISL_527813\|2020- | ···················· | ···················· | ···················· | ···················· | ···················· | ···················· |
| G | 4057 | hCoV-19/Ecuador/USFQ-199/2020\|EPI_ISL_527814\|2020- | ···················· | ···················· | ···················· | ···················· | ···················· | ···················· |
| G | 4058 | hCoV-19/Ecuador/USFQ-201/2020\|EPI_ISL_527815\|2020- | ···················· | ···················· | ···················· | ···················· | ···················· | ···················· |
| G | 4059 | hCoV-19/Ecuador/USFQ-203/2020\|EPI_ISL_527816\|2020- | ···················· | ···················· | ···················· | ···················· | ···················· | ···················· |
| G | 4060 | hCoV-19/Ecuador/USFQ-204/2020\|EPI_ISL_527817\|2020- | ···················· | ···················· | ···················· | ···················· | ···················· | ···················· |
| G | 4061 | hCoV-19/Ecuador/USFQ-206/2020\|EPI_ISL_527818\|2020- | ···················· | ···················· | ···················· | ···················· | ···················· | ···················· |
| G | 4062 | hCoV-19/Ecuador/USFQ-207/2020\|EPI_ISL_527819\|2020- | ···················· | ···················· | ···················· | ···················· | ···················· | ···················· |
| G | 4063 | hCoV-19/USA/TX-DSHS-0271/2020\|EPI_ISL_527838\|2020- | ···················· | ···················· | ···················· | ···················· | ···················· | ···················· |
| G | 4064 | hCoV-19/Brazil/SP-292/2020\|EPI_ISL_527856\|2020-04- | ···················· | ···················· | ···················· | ···················· | ···················· | ···················· |
| G | 4065 | hCoV-19/Brazil/SP-293/2020\|EPI_ISL_527857\|2020-04- | ···················· | ···················· | ···················· | ···················· | ···················· | ···················· |
| G | 4066 | hCoV-19/Brazil/SP-279/2020\|EPI_ISL_527858\|2020-03- | ···················· | ···················· | ···················· | ···················· | ···················· | ···················· |
| G | 4067 | hCoV-19/Brazil/SP-282/2020\|EPI_ISL_527859\|2020-04- | ···················· | ···················· | ···················· | ···················· | ···················· | ···················· |
| G | 4068 | hCoV-19/Brazil/SP-283/2020\|EPI_ISL_527860\|2020-04- | ···················· | ···················· | ···················· | ···················· | ···················· | ···················· |
| G | 4069 | hCoV-19/Brazil/SP-298/2020\|EPI_ISL_527861\|2020-04- | ···················· | ···················· | ···················· | ···················· | ···················· | ···················· |
| G | 4070 | hCoV-19/Brazil/SP-294/2020\|EPI_ISL_527862\|2020-03- | ···················· | ···················· | ···················· | ···················· | ···················· | ···················· |
| G | 4071 | hCoV-19/Brazil/SP-287/2020\|EPI_ISL_527863\|2020-04- | ···················· | ···················· | ···················· | ···················· | ···················· | ···················· |
| G | 4072 | hCoV-19/Brazil/SP-299/2020\|EPI_ISL_527864\|2020-04- | ···················· | ···················· | ···················· | ···················· | ···················· | ···················· |
| G | 4073 | hCoV-19/Brazil/SP-218/2020\|EPI_ISL_527865\|2020-04- | ···················· | ···················· | ···················· | ···················· | ···················· | ···················· |
| G | 4074 | hCoV-19/Brazil/SP-221/2020\|EPI_ISL_527866\|2020-04- | ···················· | ···················· | ···················· | ···················· | ···················· | ···················· |
| G | 4075 | hCoV-19/Brazil/SP-225/2020\|EPI_ISL_527867\|2020-04- | ···················· | ···················· | ···················· | ···················· | ···················· | ···················· |
| G | 4077 | hCoV-19/Brazil/SP-300/2020\|EPI_ISL_527869\|2020-04- | ···················· | ···················· | ···················· | ···················· | ···················· | ···················· |
| G | 4078 | hCoV-19/Brazil/SP-301/2020\|EPI_ISL_527870\|2020-04- | ···················· | ···················· | ···················· | ···················· | ···················· | ···················· |
| G | 4079 | hCoV-19/Nigeria/ED01-CV155/2020\|EPI_ISL_527873\|202 | ···················· | ····A··············· | ···················· | ···················· | ···················· | ···················· |
| G | 4080 | hCoV-19/Nigeria/ED02-CV156/2020\|EPI_ISL_527874\|202 | ···················· | ···················· | ···················· | ···················· | ···················· | ···················· |
| G | 4081 | hCoV-19/Nigeria/ED04-CV158/2020\|EPI_ISL_527876\|202 | ···················· | ···················· | ···················· | ···················· | ···················· | ···················· |
| G | 4082 | hCoV-19/Nigeria/ED07-CV160/2020\|EPI_ISL_527878\|202 | ···················· | ···················· | ···················· | ···················· | ···················· | ···················· |
| G | 4083 | hCoV-19/Nigeria/ED1010-CV215/2020\|EPI_ISL_527879\|2 | ···················· | ···················· | ···················· | ···················· | ···················· | ···················· |
| G | 4084 | hCoV-19/Nigeria/ED18-CV161/2020\|EPI_ISL_527880\|202 | ···················· | ···················· | ···················· | ···················· | ···················· | ···················· |
| G | 4085 | hCoV-19/Nigeria/ED20-CV162/2020\|EPI_ISL_527881\|202 | ···················· | ···················· | ···················· | ···················· | ···················· | ···················· |
| G | 4086 | hCoV-19/Nigeria/ED25-CV163/2020\|EPI_ISL_527882\|202 | ···················· | ···················· | ···················· | ···················· | ···················· | ···················· |
| G | 4087 | hCoV-19/Nigeria/ED32-CV164/2020\|EPI_ISL_527883\|202 | ···················· | ···················· | ···················· | ···················· | ···················· | ···················· |
| G | 4088 | hCoV-19/Nigeria/ED46-CV165/2020\|EPI_ISL_527884\|202 | ···················· | ···················· | ···················· | ···················· | ···················· | ···················· |
| G | 4089 | hCoV-19/Nigeria/EK291-CV115/2020\|EPI_ISL_527887\|20 | ···················· | ···················· | ···················· | ···················· | ···················· | ···················· |
| G | 4090 | hCoV-19/Nigeria/ON595-CV121/2020\|EPI_ISL_527888\|20 | ···················· | ···················· | ···················· | ···················· | ···················· | ···················· |
| G | 4091 | hCoV-19/Nigeria/ON705-CV153/2020\|EPI_ISL_527889\|20 | ···················· | ···················· | ···················· | ···················· | ···················· | ···················· |
| G | 4092 | hCoV-19/Nigeria/ON752-CV144/2020\|EPI_ISL_527890\|20 | ···················· | ···················· | ···················· | ···················· | ···················· | ···················· |
| G | 4093 | hCoV-19/Nigeria/ON920-CV152/2020\|EPI_ISL_527891\|20 | ···················· | ···················· | ···················· | ···················· | ···················· | ···················· |
| G | 4094 | hCoV-19/Nigeria/ON969-CV145/2020\|EPI_ISL_527892\|20 | ···················· | ···················· | ···················· | ···················· | ···················· | ···················· |
| G | 4095 | hCoV-19/Nigeria/OS523-CV53/2020\|EPI_ISL_527893\|202 | ···················· | ···················· | ···················· | ···················· | ···················· | ···················· |
| G | 4096 | hCoV-19/Nigeria/OY592-CV167/2020\|EPI_ISL_527898\|20 | ···················· | ···················· | ···················· | ···················· | ···················· | ···················· |
| G | 4097 | hCoV-19/Australia/VIC6071/2020\|EPI_ISL_518100\|2020 | ···················· | ···················· | ···················· | ···················· | ···················· | ···················· |
| G | 4098 | hCoV-19/Australia/VIC6072/2020\|EPI_ISL_518101\|2020 | ···················· | ···················· | ···················· | ···················· | ···················· | ···················· |
| G | 4099 | hCoV-19/Australia/VIC6074/2020\|EPI_ISL_518102\|2020 | ···················· | ···················· | ···················· | ···················· | ···················· | ···················· |
| G | 4100 | hCoV-19/Australia/VIC6077/2020\|EPI_ISL_518104\|2020 | ···················· | ···················· | ···················· | ···················· | ···················· | ···················· |
| G | 4101 | hCoV-19/Australia/VIC6078/2020\|EPI_ISL_518105\|2020 | ···················· | ···················· | ···················· | ···················· | ···················· | ···················· |
| G | 4102 | hCoV-19/Australia/VIC6079/2020\|EPI_ISL_518106\|2020 | ···················· | ···················· | ···················· | ···················· | ···················· | ···················· |
| G | 4103 | hCoV-19/Australia/VIC6080/2020\|EPI_ISL_518107\|2020 | ···················· | ···················· | ···················· | ···················· | ···················· | ···················· |
| G | 4104 | hCoV-19/Australia/VIC6082/2020\|EPI_ISL_518108\|2020 | ···················· | ···················· | ···················· | ···················· | ···················· | ···················· |
| G | 4105 | hCoV-19/Australia/VIC6083/2020\|EPI_ISL_518109\|2020 | ···················· | ···················· | ···················· | ···················· | ···················· | ···················· |
| G | 4106 | hCoV-19/Australia/VIC6084/2020\|EPI_ISL_518110\|2020 | ···················· | ···················· | ···················· | ···················· | ···················· | ···················· |
| G | 4107 | hCoV-19/Australia/VIC6085/2020\|EPI_ISL_518111\|2020 | ···················· | ···················· | ···················· | ···················· | ···················· | ···················· |
| G | 4108 | hCoV-19/Australia/VIC6087/2020\|EPI_ISL_518112\|2020 | ···················· | ···················· | ···················· | ···················· | ···················· | ···················· |
| G | 4109 | hCoV-19/Australia/VIC6088/2020\|EPI_ISL_518113\|2020 | ···················· | ···················· | ···················· | ···················· | ···················· | ···················· |
| G | 4110 | hCoV-19/Australia/VIC6089/2020\|EPI_ISL_518114\|2020 | ···················· | ···················· | ···················· | ···················· | ···················· | ···················· |
| G | 4111 | hCoV-19/Australia/VIC6090/2020\|EPI_ISL_518115\|2020 | ···················· | ···················· | ···················· | ···················· | ···················· | ···················· |
| G | 4112 | hCoV-19/Australia/VIC6091/2020\|EPI_ISL_518116\|2020 | ···················· | ···················· | ···················· | ···················· | ···················· | ···················· |
| G | 4113 | hCoV-19/Australia/VIC6093/2020\|EPI_ISL_518118\|2020 | ···················· | ···················· | ···················· | ···················· | ···················· | ···················· |
| G | 4114 | hCoV-19/Australia/VIC6094/2020\|EPI_ISL_518119\|2020 | ···················· | ···················· | ···················· | ···················· | ···················· | ···················· |
| G | 4115 | hCoV-19/Australia/VIC6095/2020\|EPI_ISL_518120\|2020 | ···················· | ···················· | ···················· | ···················· | ···················· | ···················· |
| G | 4116 | hCoV-19/Australia/VIC6096/2020\|EPI_ISL_518121\|2020 | ···················· | ···················· | ···················· | ···················· | ···················· | ···················· |
| G | 4117 | hCoV-19/Australia/VIC6098/2020\|EPI_ISL_518122\|2020 | ···················· | ···················· | ···················· | ···················· | ···················· | ···················· |
| G | 4118 | hCoV-19/Australia/VIC6099/2020\|EPI_ISL_518123\|2020 | ···················· | ···················· | ···················· | ···················· | ···················· | ···················· |
| G | 4119 | hCoV-19/Australia/VIC6101/2020\|EPI_ISL_518124\|2020 | ···················· | ···················· | ···················· | ···················· | ···················· | ···················· |
| G | 4120 | hCoV-19/Australia/VIC6102/2020\|EPI_ISL_518125\|2020 | ···················· | ···················· | ···················· | ···················· | ···················· | ···················· |
| G | 4121 | hCoV-19/Australia/VIC6106/2020\|EPI_ISL_518127\|2020 | ···················· | ···················· | ···················· | ···················· | ···················· | ···················· |
| G | 4122 | hCoV-19/Australia/VIC6107/2020\|EPI_ISL_518128\|2020 | ···················· | ···················· | ···················· | ···················· | ···················· | ···················· |
| G | 4123 | hCoV-19/Australia/VIC6109/2020\|EPI_ISL_518129\|2020 | ···················· | ···················· | ···················· | ···················· | ···················· | ···················· |
| G | 4124 | hCoV-19/Australia/VIC4856/2020\|EPI_ISL_519240\|2020 | ···················· | ···················· | ···················· | ···················· | ···················· | ···················· |
| G | 4125 | hCoV-19/Australia/VIC4866/2020\|EPI_ISL_519250\|2020 | ···················· | ···················· | ···················· | ···················· | ···················· | ···················· |
| G | 4126 | hCoV-19/Australia/VIC4867/2020\|EPI_ISL_519251\|2020 | ···················· | ···················· | ···················· | ···················· | ···················· | ···················· |
| G | 4127 | hCoV-19/Australia/VIC4868/2020\|EPI_ISL_519252\|2020 | ···················· | ···················· | ···················· | ···················· | ···················· | ···················· |
| G | 4128 | hCoV-19/Australia/VIC4869/2020\|EPI_ISL_519253\|2020 | ···················· | ···················· | ···················· | ···················· | ···················· | ···················· |
| G | 4129 | hCoV-19/Australia/VIC4870/2020\|EPI_ISL_519254\|2020 | ···················· | ···················· | ···················· | ···················· | ···················· | ···················· |
| G | 4130 | hCoV-19/Australia/VIC4871/2020\|EPI_ISL_519255\|2020 | ···················· | ···················· | ···················· | ···················· | ···················· | ···················· |
| G | 4131 | hCoV-19/Australia/VIC4874/2020\|EPI_ISL_519257\|2020 | ···················· | ···················· | ···················· | ···················· | ···················· | ···················· |
| G | 4132 | hCoV-19/Australia/VIC4875/2020\|EPI_ISL_519258\|2020 | ···················· | ···················· | ···················· | ···················· | ···················· | ···················· |
| G | 4133 | hCoV-19/Australia/VIC4876/2020\|EPI_ISL_519259\|2020 | ···················· | ···················· | ···················· | ···················· | ···················· | ···················· |
| G | 4134 | hCoV-19/Australia/VIC4877/2020\|EPI_ISL_519260\|2020 | ···················· | ···················· | ···················· | ···················· | ···················· | ···················· |
| G | 4135 | hCoV-19/Australia/VIC4879/2020\|EPI_ISL_519261\|2020 | ···················· | ···················· | ···················· | ···················· | ···················· | ···················· |

| | | | | | | | | |
|---|---|---|---|---|---|---|---|---|
| G | 4136 hCoV-19/Australia/VIC4880/2020\|EPI_ISL_519262\|2020 | .................... | .................... | .................... | .................... | .................... | .................... | |
| G | 4137 hCoV-19/Australia/VIC4881/2020\|EPI_ISL_519263\|2020 | .................... | .................... | .................... | .................... | .................... | .................... | |
| G | 4138 hCoV-19/Australia/VIC4882/2020\|EPI_ISL_519264\|2020 | .................... | .................... | .................... | .................... | .................... | .................... | |
| G | 4139 hCoV-19/Australia/VIC4883/2020\|EPI_ISL_519265\|2020 | .................... | .................... | .................... | .................... | .................... | .................... | |
| G | 4140 hCoV-19/Australia/VIC4884/2020\|EPI_ISL_519266\|2020 | .................... | .................... | .................... | .................... | .................... | .................... | |
| G | 4141 hCoV-19/Australia/VIC4885/2020\|EPI_ISL_519267\|2020 | .................... | .................... | .................... | .................... | .................... | .................... | |
| G | 4142 hCoV-19/Australia/VIC4889/2020\|EPI_ISL_519269\|2020 | .................... | .................... | .................... | .................... | .................... | .................... | |
| G | 4143 hCoV-19/Australia/VIC4890/2020\|EPI_ISL_519270\|2020 | .................... | .................... | .................... | .................... | .................... | .................... | |
| G | 4144 hCoV-19/Australia/VIC4896/2020\|EPI_ISL_519274\|2020 | .................... | .................... | .................... | .................... | .................... | .................... | |
| G | 4145 hCoV-19/Australia/VIC4897/2020\|EPI_ISL_519275\|2020 | .................... | .................... | .................... | .................... | .................... | .................... | |
| G | 4146 hCoV-19/Australia/VIC4901/2020\|EPI_ISL_519277\|2020 | .................... | .................... | .................... | .................... | .................... | .................... | |
| G | 4147 hCoV-19/Australia/VIC4903/2020\|EPI_ISL_519279\|2020 | .................... | .................... | .................... | .................... | .................... | .................... | |
| G | 4148 hCoV-19/Australia/VIC4904/2020\|EPI_ISL_519280\|2020 | .................... | .................... | .................... | .................... | .................... | .................... | |
| G | 4149 hCoV-19/Australia/VIC4905/2020\|EPI_ISL_519281\|2020 | .................... | .................... | .................... | .................... | .................... | .................... | |
| G | 4150 hCoV-19/Australia/VIC4906/2020\|EPI_ISL_519282\|2020 | .................... | .................... | .................... | .................... | .................... | .................... | |
| G | 4151 hCoV-19/Australia/VIC4907/2020\|EPI_ISL_519283\|2020 | .................... | .................... | .................... | .................... | .................... | .................... | |
| G | 4152 hCoV-19/Australia/VIC4908/2020\|EPI_ISL_519284\|2020 | .................... | .................... | .................... | .................... | .................... | .................... | |
| G | 4153 hCoV-19/Australia/VIC4909/2020\|EPI_ISL_519285\|2020 | .................... | .................... | .................... | .................... | .................... | .................... | |
| G | 4154 hCoV-19/Australia/VIC4910/2020\|EPI_ISL_519286\|2020 | .................... | .................... | .................... | .................... | .................... | .................... | |
| G | 4155 hCoV-19/Australia/VIC4911/2020\|EPI_ISL_519287\|2020 | .................... | .................... | .................... | .................... | .................... | .................... | |
| G | 4156 hCoV-19/Australia/VIC4912/2020\|EPI_ISL_519288\|2020 | .................... | .................... | .................... | .................... | .................... | .................... | |
| G | 4157 hCoV-19/Australia/VIC4913/2020\|EPI_ISL_519289\|2020 | .................... | .................... | .................... | .................... | .................... | .................... | |
| G | 4158 hCoV-19/Australia/VIC4914/2020\|EPI_ISL_519290\|2020 | .................... | .................... | .................... | .................... | .................... | .................... | |
| G | 4159 hCoV-19/Australia/VIC4915/2020\|EPI_ISL_519291\|2020 | .................... | .................... | .................... | .................... | .................... | .................... | |
| G | 4160 hCoV-19/Australia/VIC4917/2020\|EPI_ISL_519292\|2020 | .................... | .................... | .................... | .................... | .................... | .................... | |
| G | 4161 hCoV-19/Australia/VIC4918/2020\|EPI_ISL_519293\|2020 | .................... | .................... | .................... | .................... | .................... | .................... | |
| G | 4162 hCoV-19/Australia/VIC4920/2020\|EPI_ISL_519295\|2020 | .................... | .................... | .................... | .................... | .................... | .................... | |
| G | 4163 hCoV-19/Australia/VIC4921/2020\|EPI_ISL_519296\|2020 | .................... | .................... | .................... | .................... | .................... | .................... | |
| G | 4164 hCoV-19/Australia/VIC4922/2020\|EPI_ISL_519297\|2020 | .................... | .................... | .................... | .................... | .................... | .................... | |
| G | 4165 hCoV-19/Australia/VIC4924/2020\|EPI_ISL_519298\|2020 | .................... | .................... | .................... | .................... | .................... | .................... | |
| G | 4166 hCoV-19/Australia/VIC4014/2020\|EPI_ISL_519400\|2020 | .................... | .................... | .................... | .................... | .................... | .................... | |
| G | 4167 hCoV-19/Australia/VIC4015/2020\|EPI_ISL_519401\|2020 | .................... | .................... | .................... | .................... | .................... | .................... | |
| G | 4168 hCoV-19/Australia/VIC4016/2020\|EPI_ISL_519402\|2020 | .................... | .................... | .................... | .................... | .................... | .................... | |
| G | 4169 hCoV-19/Australia/VIC4017/2020\|EPI_ISL_519403\|2020 | .................... | .................... | .................... | .................... | .................... | .................... | |
| G | 4170 hCoV-19/Australia/VIC4019/2020\|EPI_ISL_519405\|2020 | .................... | .................... | .................... | .................... | .................... | .................... | |
| G | 4171 hCoV-19/Australia/VIC4020/2020\|EPI_ISL_519406\|2020 | .................... | .................... | .................... | .................... | .................... | .................... | |
| G | 4172 hCoV-19/Australia/VIC4022/2020\|EPI_ISL_519408\|2020 | .................... | .................... | .................... | .................... | .................... | .................... | |
| G | 4173 hCoV-19/Australia/VIC4023/2020\|EPI_ISL_519409\|2020 | .................... | .................... | .................... | .................... | .................... | .................... | |
| G | 4174 hCoV-19/Australia/VIC4024/2020\|EPI_ISL_519410\|2020 | .................... | .................... | .................... | .................... | .................... | .................... | |
| G | 4175 hCoV-19/Australia/VIC4031/2020\|EPI_ISL_519416\|2020 | .................... | .................... | .................... | .................... | .................... | .................... | |
| G | 4176 hCoV-19/Australia/VIC4033/2020\|EPI_ISL_519418\|2020 | .................... | .................... | .................... | .................... | .................... | .................... | |
| G | 4177 hCoV-19/Australia/VIC4039/2020\|EPI_ISL_519422\|2020 | .................... | .................... | .................... | .................... | .................... | .................... | |
| G | 4178 hCoV-19/Australia/VIC4040/2020\|EPI_ISL_519423\|2020 | .................... | .................... | .................... | .................... | .................... | .................... | |
| G | 4179 hCoV-19/Australia/VIC4041/2020\|EPI_ISL_519424\|2020 | .................... | .................... | .................... | .................... | .................... | .................... | |
| G | 4180 hCoV-19/Australia/VIC4043/2020\|EPI_ISL_519426\|2020 | .................... | .................... | .................... | .................... | .................... | .................... | |
| G | 4181 hCoV-19/Australia/VIC4046/2020\|EPI_ISL_519428\|2020 | .................... | .................... | .................... | .................... | .................... | .................... | |
| G | 4182 hCoV-19/Australia/VIC4051/2020\|EPI_ISL_519433\|2020 | .................... | .................... | .................... | .................... | .................... | .................... | |
| G | 4183 hCoV-19/Australia/VIC4054/2020\|EPI_ISL_519436\|2020 | .................... | .................... | .................... | .................... | .................... | .................... | |
| G | 4184 hCoV-19/Australia/VIC4056/2020\|EPI_ISL_519438\|2020 | .................... | .................... | .................... | .................... | .................... | .................... | |
| G | 4185 hCoV-19/Australia/VIC4057/2020\|EPI_ISL_519439\|2020 | .................... | .................... | .................... | .................... | .................... | .................... | |
| G | 4186 hCoV-19/Australia/VIC4058/2020\|EPI_ISL_519440\|2020 | .................... | .................... | .................... | .................... | .................... | .................... | |
| G | 4187 hCoV-19/Australia/VIC4060/2020\|EPI_ISL_519442\|2020 | .................... | .................... | .................... | .................... | .................... | .................... | |
| G | 4188 hCoV-19/Australia/VIC4061/2020\|EPI_ISL_519443\|2020 | .................... | .................... | .................... | .................... | .................... | .................... | |
| G | 4189 hCoV-19/Australia/VIC4064/2020\|EPI_ISL_519446\|2020 | .................... | .................... | .................... | .................... | .................... | .................... | |
| G | 4190 hCoV-19/Australia/VIC4065/2020\|EPI_ISL_519447\|2020 | .................... | .................... | .................... | .................... | .................... | .................... | |
| G | 4191 hCoV-19/Australia/VIC4067/2020\|EPI_ISL_519449\|2020 | .................... | .................... | .................... | .................... | .................... | .................... | |
| G | 4192 hCoV-19/Australia/VIC4070/2020\|EPI_ISL_519452\|2020 | .................... | .................... | .................... | .................... | .................... | .................... | |
| G | 4193 hCoV-19/Australia/VIC4071/2020\|EPI_ISL_519453\|2020 | .................... | .................... | .................... | .................... | .................... | .................... | |
| G | 4194 hCoV-19/Australia/VIC4072/2020\|EPI_ISL_519454\|2020 | .................... | .................... | .................... | .................... | .................... | .................... | |
| G | 4195 hCoV-19/Australia/VIC4077/2020\|EPI_ISL_519459\|2020 | .................... | .................... | .................... | .................... | .................... | .................... | |
| G | 4196 hCoV-19/Australia/VIC5482/2020\|EPI_ISL_520230\|2020 | .................... | .................... | .................... | .................... | .................... | .................... | |
| G | 4197 hCoV-19/Australia/VIC5486/2020\|EPI_ISL_520233\|2020 | .................... | .................... | .................... | .................... | .................... | .................... | |
| G | 4198 hCoV-19/Australia/VIC5488/2020\|EPI_ISL_520234\|2020 | .................... | .................... | .................... | .................... | .................... | .................... | |
| G | 4199 hCoV-19/Australia/VIC5489/2020\|EPI_ISL_520235\|2020 | .................... | .................... | .................... | .................... | .................... | .................... | |
| G | 4200 hCoV-19/Australia/VIC5491/2020\|EPI_ISL_520237\|2020 | .................... | .................... | .................... | .................... | .................... | .................... | |
| G | 4201 hCoV-19/Australia/VIC5492/2020\|EPI_ISL_520238\|2020 | .................... | .................... | .................... | .................... | .................... | .................... | |
| G | 4202 hCoV-19/Australia/VIC5494/2020\|EPI_ISL_520240\|2020 | .................... | .................... | .................... | .................... | .................... | .................... | |
| G | 4203 hCoV-19/Australia/VIC5495/2020\|EPI_ISL_520241\|2020 | .................... | .................... | .................... | .................... | .................... | .................... | |
| G | 4204 hCoV-19/Australia/VIC5497/2020\|EPI_ISL_520243\|2020 | .................... | .................... | .................... | .................... | .................... | .................... | |
| G | 4205 hCoV-19/Australia/VIC5498/2020\|EPI_ISL_520244\|2020 | .................... | .................... | .................... | .................... | .................... | .................... | |
| G | 4206 hCoV-19/Australia/VIC5499/2020\|EPI_ISL_520245\|2020 | .................... | .................... | .................... | .................... | .................... | .................... | |
| G | 4207 hCoV-19/Australia/VIC5500/2020\|EPI_ISL_520246\|2020 | .................... | .................... | .................... | .................... | .................... | .................... | |
| G | 4208 hCoV-19/Australia/VIC5501/2020\|EPI_ISL_520247\|2020 | .................... | .................... | .................... | .................... | .................... | .................... | |
| G | 4209 hCoV-19/Australia/VIC5502/2020\|EPI_ISL_520248\|2020 | .................... | .................... | .................... | .................... | .................... | .................... | |
| G | 4211 hCoV-19/Australia/VIC3494/2020\|EPI_ISL_520250\|2020 | .................... | .................... | .................... | .................... | .NNN................ | .................... | 1 |
| G | 4212 hCoV-19/Australia/VIC3495/2020\|EPI_ISL_520251\|2020 | .................... | .................... | .................... | .................... | .................... | .................... | |
| G | 4213 hCoV-19/Australia/VIC3496/2020\|EPI_ISL_520252\|2020 | .................... | .................... | .................... | .................... | .................... | .................... | |
| G | 4214 hCoV-19/Australia/VIC3498/2020\|EPI_ISL_520254\|2020 | .................... | .................... | .................... | .................... | .................... | .................... | |
| G | 4215 hCoV-19/Australia/VIC3499/2020\|EPI_ISL_520255\|2020 | .................... | .................... | .................... | .................... | .................... | .................... | |
| G | 4216 hCoV-19/Australia/VIC3500/2020\|EPI_ISL_520256\|2020 | .................... | .................... | .................... | .................... | .................... | .................... | |

| | | | | | | | | | |
|---|---|---|---|---|---|---|---|---|---|
| G | 4218 | hCoV-19/Australia/VIC3504/2020\|EPI_ISL_520259\|2020 | .................... | .................... | .................... | .................... | .................... | .................... | |
| G | 4219 | hCoV-19/Australia/VIC3505/2020\|EPI_ISL_520260\|2020 | .................... | .................... | .................... | .................... | .................... | .................... | |
| G | 4220 | hCoV-19/Australia/VIC3506/2020\|EPI_ISL_520261\|2020 | .................... | .................... | .................... | .................... | .................... | .................... | |
| G | 4221 | hCoV-19/Australia/VIC3507/2020\|EPI_ISL_520262\|2020 | .................... | .................... | .................... | .................... | .................... | .................... | |
| G | 4222 | hCoV-19/Australia/VIC3508/2020\|EPI_ISL_520263\|2020 | .................... | .................... | .................... | .................... | .................... | .................... | |
| G | 4223 | hCoV-19/Australia/VIC3509/2020\|EPI_ISL_520264\|2020 | .................... | .................... | .................... | .................... | .................... | .................... | |
| G | 4224 | hCoV-19/Australia/VIC3511/2020\|EPI_ISL_520265\|2020 | .................... | .................... | .................... | .................... | .................... | .................... | |
| G | 4225 | hCoV-19/Australia/VIC3514/2020\|EPI_ISL_520267\|2020 | .................... | .................... | .................... | .................... | .................... | .................... | |
| G | 4226 | hCoV-19/Australia/VIC3516/2020\|EPI_ISL_520268\|2020 | .................... | .................... | .................... | .................... | .................... | .................... | |
| G | 4227 | hCoV-19/Australia/VIC3520/2020\|EPI_ISL_520272\|2020 | .................... | .................... | .................... | .................... | .................... | .................... | |
| G | 4228 | hCoV-19/Australia/VIC3521/2020\|EPI_ISL_520273\|2020 | .................... | .................... | .................... | .................... | .................... | .................... | |
| G | 4229 | hCoV-19/Australia/VIC3522/2020\|EPI_ISL_520274\|2020 | .................... | .................... | .................... | .................... | .................... | .................... | |
| G | 4230 | hCoV-19/Australia/VIC3524/2020\|EPI_ISL_520275\|2020 | .................... | .................... | .................... | .................... | .................... | .................... | |
| G | 4232 | hCoV-19/Australia/VIC3528/2020\|EPI_ISL_520277\|2020 | .................... | .................... | .................... | .................... | .................... | .................... | |
| G | 4233 | hCoV-19/Australia/VIC3529/2020\|EPI_ISL_520278\|2020 | .................... | .................... | .................... | .................... | .................... | .................... | |
| G | 4234 | hCoV-19/Australia/VIC3530/2020\|EPI_ISL_520279\|2020 | .................... | .................... | .................... | .................... | .................... | .................... | |
| G | 4235 | hCoV-19/Australia/VIC3531/2020\|EPI_ISL_520280\|2020 | .................... | .................... | .................... | .................... | .................... | .................... | |
| G | 4236 | hCoV-19/Australia/VIC3532/2020\|EPI_ISL_520281\|2020 | .................... | .................... | .................... | .................... | .................... | .................... | |
| G | 4237 | hCoV-19/Australia/VIC3533/2020\|EPI_ISL_520282\|2020 | .................... | .................... | .................... | .................... | .................... | .................... | |
| G | 4239 | hCoV-19/Australia/VIC3535/2020\|EPI_ISL_520284\|2020 | .................... | .................... | .................... | .................... | .................... | .................... | |
| G | 4241 | hCoV-19/Australia/VIC3540/2020\|EPI_ISL_520289\|2020 | .................... | .................... | .................... | .................... | ..N................. | .................... | |
| G | 4242 | hCoV-19/Australia/VIC3684/2020\|EPI_ISL_520421\|2020 | .................... | .................... | .................... | .................... | .................... | .................... | |
| G | 4243 | hCoV-19/Australia/VIC2723/2020\|EPI_ISL_521560\|2020 | .................... | .................... | .................... | .................... | .................... | .................... | |
| G | 4244 | hCoV-19/Australia/VIC2724/2020\|EPI_ISL_521561\|2020 | .................... | .................... | .................... | .................... | .................... | .................... | |
| G | 4245 | hCoV-19/Australia/VIC2727/2020\|EPI_ISL_521562\|2020 | .................... | .................... | .................... | .................... | .................... | .................... | |
| G | 4246 | hCoV-19/Australia/VIC2728/2020\|EPI_ISL_521563\|2020 | .................... | .................... | .................... | .................... | .................... | .................... | |
| G | 4247 | hCoV-19/Australia/VIC2729/2020\|EPI_ISL_521564\|2020 | .................... | .................... | .................... | .................... | .................... | .................... | |
| G | 4248 | hCoV-19/Australia/VIC2730/2020\|EPI_ISL_521565\|2020 | .................... | .................... | .................... | .................... | .................... | .................... | |
| G | 4249 | hCoV-19/Australia/VIC2731/2020\|EPI_ISL_521566\|2020 | .................... | .................... | .................... | .................... | .................... | .................... | |
| G | 4250 | hCoV-19/Australia/VIC2732/2020\|EPI_ISL_521567\|2020 | .................... | .................... | .................... | .................... | .................... | .................... | |
| G | 4251 | hCoV-19/Australia/VIC2733/2020\|EPI_ISL_521568\|2020 | .................... | .................... | .................... | .................... | .................... | .................... | |
| G | 4252 | hCoV-19/Australia/VIC2734/2020\|EPI_ISL_521569\|2020 | .................... | .................... | .................... | .................... | .................... | .................... | |
| G | 4253 | hCoV-19/Australia/VIC2735/2020\|EPI_ISL_521570\|2020 | .................... | .................... | .................... | .................... | .................... | .................... | |
| G | 4254 | hCoV-19/Australia/VIC2736/2020\|EPI_ISL_521571\|2020 | .................... | .................... | .................... | .................... | .................... | .................... | |
| G | 4255 | hCoV-19/Australia/VIC2738/2020\|EPI_ISL_521573\|2020 | .................... | .................... | .................... | .................... | .................... | .................... | |
| G | 4256 | hCoV-19/Australia/VIC2739/2020\|EPI_ISL_521574\|2020 | .................... | .................... | .................... | .................... | .................... | .................... | |
| G | 4257 | hCoV-19/Australia/VIC2740/2020\|EPI_ISL_521575\|2020 | .................... | .................... | .................... | .................... | .................... | .................... | |
| G | 4258 | hCoV-19/Australia/VIC2741/2020\|EPI_ISL_521576\|2020 | .................... | .................... | .................... | .................... | .................... | .................... | |
| G | 4259 | hCoV-19/Australia/VIC2742/2020\|EPI_ISL_521577\|2020 | .................... | .................... | .................... | .................... | .................... | .................... | |
| G | 4260 | hCoV-19/Australia/VIC2743/2020\|EPI_ISL_521578\|2020 | .................... | .................... | .................... | .................... | .................... | .................... | |
| G | 4261 | hCoV-19/Australia/VIC2744/2020\|EPI_ISL_521579\|2020 | .................... | .................... | .................... | .................... | .................... | .................... | |
| G | 4262 | hCoV-19/Australia/VIC2745/2020\|EPI_ISL_521580\|2020 | .................... | .................... | .................... | .................... | .................... | .................... | |
| G | 4263 | hCoV-19/Australia/VIC2746/2020\|EPI_ISL_521581\|2020 | .................... | .................... | .................... | .................... | .................... | .................... | |
| G | 4264 | hCoV-19/Australia/VIC2747/2020\|EPI_ISL_521582\|2020 | .................... | .................... | .................... | .................... | .................... | .................... | |
| G | 4265 | hCoV-19/Australia/VIC2748/2020\|EPI_ISL_521583\|2020 | .................... | .................... | .................... | .................... | .................... | .................... | |
| G | 4266 | hCoV-19/Australia/VIC2749/2020\|EPI_ISL_521584\|2020 | .................... | .................... | .................... | .................... | .................... | .................... | |
| G | 4267 | hCoV-19/Australia/VIC2750/2020\|EPI_ISL_521585\|2020 | .................... | .................... | .................... | .................... | .................... | .................... | |
| G | 4268 | hCoV-19/Australia/VIC2751/2020\|EPI_ISL_521586\|2020 | .................... | .................... | .................... | .................... | .................... | .................... | |
| G | 4270 | hCoV-19/Australia/VIC2753/2020\|EPI_ISL_521588\|2020 | .................... | .................... | .................... | .................... | NNN................. | .................... | 1 |
| G | 4271 | hCoV-19/Australia/VIC2754/2020\|EPI_ISL_521589\|2020 | .................... | .................... | .................... | .................... | .................... | .................... | |
| G | 4272 | hCoV-19/Australia/VIC2755/2020\|EPI_ISL_521590\|2020 | .................... | .................... | .................... | .................... | .................... | .................... | |
| G | 4273 | hCoV-19/Australia/VIC2756/2020\|EPI_ISL_521591\|2020 | .................... | .................... | .................... | .................... | .................... | .................... | |
| G | 4274 | hCoV-19/Australia/VIC2757/2020\|EPI_ISL_521592\|2020 | .................... | .................... | .................... | .................... | .................... | .................... | |
| G | 4275 | hCoV-19/Australia/VIC2758/2020\|EPI_ISL_521593\|2020 | .................... | .................... | .................... | .................... | .................... | .................... | |
| G | 4276 | hCoV-19/Australia/VIC2759/2020\|EPI_ISL_521594\|2020 | .................... | .................... | .................... | .................... | .................... | .................... | |
| G | 4277 | hCoV-19/Australia/VIC2761/2020\|EPI_ISL_521596\|2020 | .................... | .................... | .................... | .................... | .................... | .................... | |
| G | 4280 | hCoV-19/Australia/VIC2869/2020\|EPI_ISL_521700\|2020 | .................... | .................... | .................... | .................... | .................... | .................... | |
| G | 4281 | hCoV-19/Australia/VIC2870/2020\|EPI_ISL_521701\|2020 | .................... | .................... | .................... | .................... | .................... | .................... | |
| G | 4282 | hCoV-19/Australia/VIC2871/2020\|EPI_ISL_521702\|2020 | .................... | .................... | .................... | .................... | .................... | .................... | |
| G | 4283 | hCoV-19/Australia/VIC2872/2020\|EPI_ISL_521703\|2020 | .................... | .................... | .................... | .................... | .................... | .................... | |
| G | 4284 | hCoV-19/Australia/VIC2873/2020\|EPI_ISL_521704\|2020 | .................... | .................... | .................... | .................... | .................... | .................... | |
| G | 4285 | hCoV-19/Australia/VIC2874/2020\|EPI_ISL_521705\|2020 | .................... | .................... | .................... | .................... | .................... | .................... | |
| G | 4286 | hCoV-19/Australia/VIC2875/2020\|EPI_ISL_521706\|2020 | .................... | .................... | .................... | .................... | .................... | .................... | |
| G | 4287 | hCoV-19/Australia/VIC2876/2020\|EPI_ISL_521707\|2020 | .................... | .................... | .................... | .................... | .................... | .................... | |
| G | 4288 | hCoV-19/Australia/VIC2877/2020\|EPI_ISL_521708\|2020 | .................... | .................... | .................... | .................... | .................... | .................... | |
| G | 4289 | hCoV-19/Australia/VIC2878/2020\|EPI_ISL_521709\|2020 | .................... | .................... | .................... | .................... | .NNN................ | .................... | 1 |
| G | 4290 | hCoV-19/Australia/VIC2879/2020\|EPI_ISL_521710\|2020 | .................... | .................... | .................... | .................... | .................... | .................... | |
| G | 4291 | hCoV-19/Australia/VIC2880/2020\|EPI_ISL_521711\|2020 | .................... | .................... | .................... | .................... | .................... | .................... | |
| G | 4292 | hCoV-19/Australia/VIC2881/2020\|EPI_ISL_521712\|2020 | .................... | .................... | .................... | .................... | .................... | .................... | |
| G | 4293 | hCoV-19/Australia/VIC2882/2020\|EPI_ISL_521713\|2020 | .................... | .................... | .................... | .................... | .................... | .................... | |
| G | 4294 | hCoV-19/Australia/VIC2883/2020\|EPI_ISL_521714\|2020 | .................... | .................... | .................... | .................... | .................... | .................... | |
| G | 4295 | hCoV-19/Australia/VIC2884/2020\|EPI_ISL_521715\|2020 | .................... | .................... | .................... | .................... | .................... | .................... | |
| G | 4296 | hCoV-19/Australia/VIC2885/2020\|EPI_ISL_521716\|2020 | .................... | .................... | .................... | .................... | .................... | .................... | |
| G | 4297 | hCoV-19/Australia/VIC2886/2020\|EPI_ISL_521717\|2020 | .................... | .................... | .................... | .................... | .................... | .................... | |
| G | 4298 | hCoV-19/Australia/VIC2887/2020\|EPI_ISL_521718\|2020 | .................... | .................... | .................... | .................... | .................... | .................... | |
| G | 4299 | hCoV-19/Australia/VIC2888/2020\|EPI_ISL_521719\|2020 | .................... | .................... | .................... | .................... | .................... | .................... | |
| G | 4300 | hCoV-19/Australia/VIC2889/2020\|EPI_ISL_521720\|2020 | .................... | .................... | .................... | .................... | .................... | .................... | |
| G | 4301 | hCoV-19/Australia/VIC2890/2020\|EPI_ISL_521721\|2020 | .................... | .................... | .................... | .................... | .................... | .................... | |
| G | 4302 | hCoV-19/Australia/VIC2891/2020\|EPI_ISL_521722\|2020 | .................... | .................... | .................... | .................... | .................... | .................... | |
| G | 4303 | hCoV-19/Australia/VIC2892/2020\|EPI_ISL_521723\|2020 | .................... | .................... | .................... | .................... | .................... | .................... | |

| | | | | | | | | |
|---|---|---|---|---|---|---|---|---|
| G | 4304 | hCoV-19/Australia/VIC2893/2020\|EPI_ISL_521724\|2020 | .................... | .................... | ..................... | .................... | .................... | .................... |
| G | 4305 | hCoV-19/Australia/VIC2894/2020\|EPI_ISL_521725\|2020 | .................... | .................... | ..................... | .................... | .................... | .................... |
| G | 4306 | hCoV-19/Australia/VIC2895/2020\|EPI_ISL_521726\|2020 | .................... | .................... | ..................... | .................... | .................... | .................... |
| G | 4307 | hCoV-19/Australia/VIC2896/2020\|EPI_ISL_521727\|2020 | .................... | .................... | ..................... | .................... | .................... | .................... |
| G | 4308 | hCoV-19/Australia/VIC2898/2020\|EPI_ISL_521729\|2020 | .................... | .................... | ..................... | .................... | .................... | .................... |
| G | 4309 | hCoV-19/Australia/VIC2899/2020\|EPI_ISL_521730\|2020 | .................... | .................... | ..................... | .................... | .................... | .................... |
| G | 4310 | hCoV-19/Australia/VIC2900/2020\|EPI_ISL_521731\|2020 | .................... | .................... | ..................... | .................... | .................... | .................... |
| G | 4311 | hCoV-19/Australia/VIC2901/2020\|EPI_ISL_521732\|2020 | .................... | .................... | ..................... | .................... | .................... | .................... |
| G | 4312 | hCoV-19/Australia/VIC2902/2020\|EPI_ISL_521733\|2020 | .................... | .................... | ..................... | .................... | .................... | .................... |
| G | 4313 | hCoV-19/Australia/VIC2903/2020\|EPI_ISL_521734\|2020 | .................... | .................... | ..................... | .................... | .................... | .................... |
| G | 4314 | hCoV-19/Australia/VIC2905/2020\|EPI_ISL_521736\|2020 | .................... | .................... | ..................... | .................... | .................... | .................... |
| G | 4315 | hCoV-19/Australia/VIC2906/2020\|EPI_ISL_521737\|2020 | .................... | .................... | ..................... | .................... | .................... | .................... |
| G | 4316 | hCoV-19/Australia/VIC2907/2020\|EPI_ISL_521738\|2020 | .................... | .................... | ..................... | .................... | .................... | .................... |
| G | 4317 | hCoV-19/Australia/VIC2908/2020\|EPI_ISL_521739\|2020 | .................... | .................... | ..................... | .................... | .................... | .................... |
| G | 4318 | hCoV-19/Australia/VIC2909/2020\|EPI_ISL_521740\|2020 | .................... | .................... | ..................... | .................... | .................... | .................... |
| G | 4319 | hCoV-19/Australia/VIC2910/2020\|EPI_ISL_521741\|2020 | .................... | .................... | ..................... | .................... | .................... | .................... |
| G | 4320 | hCoV-19/Australia/VIC2911/2020\|EPI_ISL_521742\|2020 | .................... | .................... | ..................... | .................... | .................... | .................... |
| G | 4321 | hCoV-19/Australia/VIC2912/2020\|EPI_ISL_521743\|2020 | .................... | .................... | ..................... | .................... | .................... | .................... |
| G | 4322 | hCoV-19/Australia/VIC2913/2020\|EPI_ISL_521744\|2020 | .................... | .................... | ..................... | .................... | .................... | .................... |
| G | 4323 | hCoV-19/Australia/VIC2914/2020\|EPI_ISL_521745\|2020 | .................... | .................... | ..................... | .................... | .................... | .................... |
| G | 4324 | hCoV-19/Australia/VIC2915/2020\|EPI_ISL_521746\|2020 | .................... | .................... | ..................... | .................... | .................... | .................... |
| G | 4325 | hCoV-19/Australia/VIC2916/2020\|EPI_ISL_521747\|2020 | .................... | .................... | ..................... | .................... | .................... | .................... |
| G | 4326 | hCoV-19/Australia/VIC2917/2020\|EPI_ISL_521748\|2020 | .................... | .................... | ..................... | .................... | .................... | .................... |
| G | 4327 | hCoV-19/Australia/VIC2918/2020\|EPI_ISL_521749\|2020 | .................... | .................... | ..................... | .................... | .................... | .................... |
| G | 4328 | hCoV-19/Australia/VIC2919/2020\|EPI_ISL_521750\|2020 | .................... | .................... | ..................... | .................... | .................... | .................... |
| G | 4329 | hCoV-19/Australia/VIC2920/2020\|EPI_ISL_521751\|2020 | .................... | .................... | ..................... | .................... | .................... | .................... |
| G | 4330 | hCoV-19/Australia/VIC2921/2020\|EPI_ISL_521752\|2020 | .................... | .................... | ..................... | .................... | .................... | .................... |
| G | 4331 | hCoV-19/Australia/VIC2922/2020\|EPI_ISL_521753\|2020 | .................... | .................... | ..................... | .................... | .................... | .................... |
| G | 4332 | hCoV-19/Australia/VIC2923/2020\|EPI_ISL_521754\|2020 | .................... | .................... | ..................... | .................... | .................... | .................... |
| G | 4333 | hCoV-19/Australia/VIC2924/2020\|EPI_ISL_521755\|2020 | .................... | .................... | ..................... | .................... | .................... | .................... |
| G | 4334 | hCoV-19/Australia/VIC2925/2020\|EPI_ISL_521756\|2020 | .................... | .................... | ..................... | .................... | .................... | .................... |
| G | 4335 | hCoV-19/Australia/VIC2926/2020\|EPI_ISL_521757\|2020 | .................... | .................... | ..................... | .................... | .................... | .................... |
| G | 4337 | hCoV-19/Australia/VIC2929/2020\|EPI_ISL_521760\|2020 | .................... | .................... | ..................... | .................... | .................... | .................... |
| G | 4338 | hCoV-19/Australia/VIC2930/2020\|EPI_ISL_521761\|2020 | .................... | .................... | ..................... | .................... | .................... | .................... |
| G | 4339 | hCoV-19/Australia/VIC2931/2020\|EPI_ISL_521762\|2020 | .................... | .................... | ..................... | .................... | .................... | .................... |
| G | 4340 | hCoV-19/Australia/VIC2932/2020\|EPI_ISL_521763\|2020 | .................... | .................... | ..................... | .................... | .................... | .................... |
| G | 4341 | hCoV-19/Australia/VIC2933/2020\|EPI_ISL_521764\|2020 | .................... | .................... | ..................... | .................... | .................... | .................... |
| G | 4343 | hCoV-19/Australia/VIC2935/2020\|EPI_ISL_521766\|2020 | .................... | .................... | ..................... | .................... | .................... | .................... |
| G | 4344 | hCoV-19/Australia/VIC2938/2020\|EPI_ISL_521769\|2020 | .................... | .................... | ..................... | .................... | .................... | .................... |
| G | 4345 | hCoV-19/USA/WI-UW-1133/2020\|EPI_ISL_522890\|2020-08 | .................... | .................... | ..................... | .................... | .................... | .................... |
| G | 4346 | hCoV-19/USA/WI-UW-1134/2020\|EPI_ISL_522891\|2020-08 | .................... | .................... | ..................... | .................... | .................... | .................... |
| G | 4347 | hCoV-19/USA/WI-UW-1135/2020\|EPI_ISL_522892\|2020-08 | .................... | .................... | ..................... | .................... | .................... | .................... |
| G | 4349 | hCoV-19/USA/WI-UW-1138/2020\|EPI_ISL_522895\|2020-08 | .................... | .................... | ..................... | .................... | .................... | .................... |
| G | 4350 | hCoV-19/USA/WI-UW-1139/2020\|EPI_ISL_522896\|2020-08 | .................... | .................... | ..................... | .................... | .................... | .................... |
| G | 4351 | hCoV-19/USA/WI-UW-1141/2020\|EPI_ISL_522898\|2020-08 | .................... | .................... | ..................... | .................... | .................... | .................... |
| G | 4352 | hCoV-19/USA/WI-UW-1142/2020\|EPI_ISL_522899\|2020-08 | .................... | .................... | ..................... | .................... | .................... | .................... |
| G | 4353 | hCoV-19/Australia/VIC4836/2020\|EPI_ISL_519220\|2020 | .................... | .................... | ..................... | .................... | .................... | .................... |
| G | 4354 | hCoV-19/Australia/VIC4837/2020\|EPI_ISL_519221\|2020 | .................... | .................... | ..................... | .................... | .................... | .................... |
| G | 4355 | hCoV-19/Australia/VIC4838/2020\|EPI_ISL_519222\|2020 | .................... | .................... | ..................... | .................... | .................... | .................... |
| G | 4356 | hCoV-19/Australia/VIC4839/2020\|EPI_ISL_519223\|2020 | .................... | .................... | ..................... | .................... | .................... | .................... |
| G | 4357 | hCoV-19/Australia/VIC4840/2020\|EPI_ISL_519224\|2020 | .................... | .................... | ..................... | .................... | .................... | .................... |
| G | 4358 | hCoV-19/Australia/VIC4841/2020\|EPI_ISL_519225\|2020 | .................... | .................... | ..................... | .................... | .................... | .................... |
| G | 4359 | hCoV-19/Australia/VIC4842/2020\|EPI_ISL_519226\|2020 | .................... | .................... | ..................... | .................... | .................... | .................... |
| G | 4360 | hCoV-19/Australia/VIC4844/2020\|EPI_ISL_519228\|2020 | .................... | .................... | ..................... | .................... | .................... | .................... |
| G | 4361 | hCoV-19/Australia/VIC4845/2020\|EPI_ISL_519229\|2020 | .................... | .................... | ..................... | .................... | .................... | .................... |
| G | 4362 | hCoV-19/Australia/VIC4847/2020\|EPI_ISL_519231\|2020 | .................... | .................... | ..................... | .................... | .................... | .................... |
| G | 4363 | hCoV-19/Australia/VIC4848/2020\|EPI_ISL_519232\|2020 | .................... | .................... | ..................... | .................... | .................... | .................... |
| G | 4364 | hCoV-19/Australia/VIC4849/2020\|EPI_ISL_519233\|2020 | .................... | .................... | ..................... | .................... | .................... | .................... |
| G | 4365 | hCoV-19/Australia/VIC4850/2020\|EPI_ISL_519234\|2020 | .................... | .................... | ..................... | .................... | .................... | .................... |
| G | 4366 | hCoV-19/Australia/VIC4851/2020\|EPI_ISL_519235\|2020 | .................... | .................... | ..................... | .................... | .................... | .................... |
| G | 4367 | hCoV-19/Australia/VIC4852/2020\|EPI_ISL_519236\|2020 | .................... | .................... | ..................... | .................... | .................... | .................... |
| G | 4368 | hCoV-19/Australia/VIC4853/2020\|EPI_ISL_519237\|2020 | .................... | .................... | ..................... | .................... | .................... | .................... |
| G | 4369 | hCoV-19/Australia/VIC4854/2020\|EPI_ISL_519238\|2020 | .................... | .................... | ..................... | .................... | .................... | .................... |
| G | 4370 | hCoV-19/Australia/VIC4855/2020\|EPI_ISL_519239\|2020 | .................... | .................... | ..................... | .................... | .................... | .................... |
| G | 4371 | hCoV-19/Australia/VIC4857/2020\|EPI_ISL_519241\|2020 | .................... | .................... | ..................... | .................... | .................... | .................... |
| G | 4372 | hCoV-19/Australia/VIC4858/2020\|EPI_ISL_519242\|2020 | .................... | .................... | ..................... | .................... | .................... | .................... |
| G | 4373 | hCoV-19/Australia/VIC4859/2020\|EPI_ISL_519243\|2020 | .................... | .................... | ..................... | .................... | .................... | .................... |
| G | 4374 | hCoV-19/Australia/VIC4860/2020\|EPI_ISL_519244\|2020 | .................... | .................... | ..................... | .................... | .................... | .................... |
| G | 4375 | hCoV-19/Australia/VIC4861/2020\|EPI_ISL_519245\|2020 | .................... | .................... | ..................... | .................... | .................... | .................... |
| G | 4376 | hCoV-19/Australia/VIC4862/2020\|EPI_ISL_519246\|2020 | .................... | .................... | ..................... | .................... | .................... | .................... |
| G | 4377 | hCoV-19/Australia/VIC4863/2020\|EPI_ISL_519247\|2020 | .................... | .................... | ..................... | .................... | .................... | .................... |
| G | 4378 | hCoV-19/Australia/VIC4864/2020\|EPI_ISL_519248\|2020 | .................... | .................... | ..................... | .................... | .................... | .................... |
| G | 4379 | hCoV-19/Australia/VIC4865/2020\|EPI_ISL_519249\|2020 | .................... | .................... | ..................... | .................... | .................... | .................... |
| G | 4380 | hCoV-19/Australia/VIC5453/2020\|EPI_ISL_520203\|2020 | .................... | .................... | ..................... | .................... | .................... | .................... |
| G | 4381 | hCoV-19/Australia/VIC5454/2020\|EPI_ISL_520204\|2020 | .................... | .................... | ..................... | .................... | .................... | .................... |
| G | 4382 | hCoV-19/Australia/VIC5455/2020\|EPI_ISL_520205\|2020 | .................... | .................... | ..................... | .................... | .................... | .................... |
| G | 4383 | hCoV-19/Australia/VIC5456/2020\|EPI_ISL_520206\|2020 | .................... | .................... | ..................... | .................... | .................... | .................... |
| G | 4384 | hCoV-19/Australia/VIC5457/2020\|EPI_ISL_520207\|2020 | .................... | .................... | ..................... | .................... | .................... | .................... |
| G | 4385 | hCoV-19/Australia/VIC5458/2020\|EPI_ISL_520208\|2020 | .................... | .................... | ..................... | .................... | .................... | .................... |
| G | 4386 | hCoV-19/Australia/VIC5459/2020\|EPI_ISL_520209\|2020 | .................... | .................... | ..................... | .................... | .................... | .................... |

| | | | | | | | | |
|---|---|---|---|---|---|---|---|---|
| G | 4387 hCoV-19/Australia/VIC5460/2020\|EPI_ISL_520210\|2020 | ···················· | ···················· | ····················· | ···················· | ···················· | ···················· | |
| G | 4388 hCoV-19/Australia/VIC5461/2020\|EPI_ISL_520211\|2020 | ···················· | ···················· | ····················· | ···················· | ···················· | ···················· | |
| G | 4389 hCoV-19/Australia/VIC5462/2020\|EPI_ISL_520212\|2020 | ···················· | ···················· | ····················· | ···················· | ···················· | ···················· | |
| G | 4390 hCoV-19/Australia/VIC5464/2020\|EPI_ISL_520213\|2020 | ···················· | ···················· | ····················· | ···················· | ···················· | ···················· | |
| G | 4391 hCoV-19/Australia/VIC5465/2020\|EPI_ISL_520214\|2020 | ···················· | ···················· | ····················· | ···················· | ···················· | ···················· | |
| G | 4392 hCoV-19/Australia/VIC5466/2020\|EPI_ISL_520215\|2020 | ···················· | ···················· | ····················· | ···················· | ···················· | ···················· | |
| G | 4393 hCoV-19/Australia/VIC5467/2020\|EPI_ISL_520216\|2020 | ···················· | ···················· | ····················· | ···················· | ···················· | ···················· | |
| G | 4394 hCoV-19/Australia/VIC5468/2020\|EPI_ISL_520217\|2020 | ···················· | ···················· | ····················· | ···················· | ···················· | ···················· | |
| G | 4395 hCoV-19/Australia/VIC5469/2020\|EPI_ISL_520218\|2020 | ···················· | ···················· | ····················· | ···················· | ···················· | ···················· | |
| G | 4396 hCoV-19/Australia/VIC5470/2020\|EPI_ISL_520219\|2020 | ···················· | ···················· | ····················· | ···················· | ···················· | ···················· | |
| G | 4397 hCoV-19/Australia/VIC5471/2020\|EPI_ISL_520220\|2020 | ···················· | ···················· | ····················· | ···················· | ···················· | ···················· | |
| G | 4398 hCoV-19/Australia/VIC5472/2020\|EPI_ISL_520221\|2020 | ···················· | ···················· | ····················· | ···················· | ···················· | ···················· | |
| G | 4399 hCoV-19/Australia/VIC5473/2020\|EPI_ISL_520222\|2020 | ···················· | ···················· | ····················· | ···················· | ···················· | ···················· | |
| G | 4400 hCoV-19/Australia/VIC5474/2020\|EPI_ISL_520223\|2020 | ···················· | ···················· | ····················· | ···················· | ···················· | ···················· | |
| G | 4401 hCoV-19/Australia/VIC5475/2020\|EPI_ISL_520224\|2020 | ···················· | ···················· | ····················· | ···················· | ···················· | ···················· | |
| G | 4402 hCoV-19/Australia/VIC5477/2020\|EPI_ISL_520225\|2020 | ···················· | ···················· | ····················· | ···················· | ···················· | ···················· | |
| G | 4403 hCoV-19/Australia/VIC5478/2020\|EPI_ISL_520226\|2020 | ···················· | ···················· | ····················· | ···················· | ···················· | ···················· | |
| G | 4404 hCoV-19/Australia/VIC5479/2020\|EPI_ISL_520227\|2020 | ···················· | ···················· | ····················· | ···················· | ···················· | ···················· | |
| G | 4405 hCoV-19/Australia/VIC5480/2020\|EPI_ISL_520228\|2020 | ···················· | ···················· | ····················· | ···················· | ···················· | ···················· | |
| G | 4406 hCoV-19/Australia/VIC5481/2020\|EPI_ISL_520229\|2020 | ···················· | ···················· | ····················· | ···················· | ···················· | ···················· | |
| G | 4407 hCoV-19/Australia/VIC3541/2020\|EPI_ISL_520290\|2020 | ···················· | ···················· | ····················· | ···················· | ···················· | ···················· | |
| G | 4408 hCoV-19/Australia/VIC3542/2020\|EPI_ISL_520291\|2020 | ···················· | ···················· | ····················· | ···················· | ···················· | ···················· | |
| G | 4410 hCoV-19/Australia/VIC3544/2020\|EPI_ISL_520293\|2020 | ···················· | ···················· | ····················· | ···················· | ···················· | ···················· | |
| G | 4411 hCoV-19/Australia/VIC3545/2020\|EPI_ISL_520294\|2020 | ···················· | ···················· | ····················· | ···················· | ···················· | ···················· | |
| G | 4412 hCoV-19/Australia/VIC3546/2020\|EPI_ISL_520295\|2020 | ···················· | ···················· | ····················· | ···················· | NNN················· | ···················· | 1 |
| G | 4413 hCoV-19/Australia/VIC3548/2020\|EPI_ISL_520297\|2020 | ···················· | ···················· | ····················· | ···················· | ···················· | ···················· | |
| G | 4414 hCoV-19/Australia/VIC2694/2020\|EPI_ISL_521534\|2020 | ···················· | ···················· | ····················· | ···················· | ···················· | ···················· | |
| G | 4415 hCoV-19/Australia/VIC2695/2020\|EPI_ISL_521535\|2020 | ···················· | ···················· | ····················· | ···················· | ···················· | ···················· | |
| G | 4416 hCoV-19/Australia/VIC2696/2020\|EPI_ISL_521536\|2020 | ···················· | ···················· | ····················· | ···················· | ···················· | ···················· | |
| G | 4417 hCoV-19/Australia/VIC2698/2020\|EPI_ISL_521537\|2020 | ···················· | ···················· | ····················· | ···················· | ···················· | ···················· | |
| G | 4418 hCoV-19/Australia/VIC2699/2020\|EPI_ISL_521538\|2020 | ···················· | ···················· | ····················· | ···················· | ···················· | ···················· | |
| G | 4419 hCoV-19/Australia/VIC2700/2020\|EPI_ISL_521539\|2020 | ···················· | ···················· | ····················· | ···················· | ···················· | ···················· | |
| G | 4420 hCoV-19/Australia/VIC2701/2020\|EPI_ISL_521540\|2020 | ···················· | ···················· | ····················· | ···················· | ···················· | ···················· | |
| G | 4421 hCoV-19/Australia/VIC2703/2020\|EPI_ISL_521541\|2020 | ···················· | ···················· | ····················· | ···················· | ···················· | ···················· | |
| G | 4422 hCoV-19/Australia/VIC2705/2020\|EPI_ISL_521543\|2020 | ···················· | ···················· | ····················· | ···················· | ···················· | ···················· | |
| G | 4423 hCoV-19/Australia/VIC2707/2020\|EPI_ISL_521545\|2020 | ···················· | ···················· | ····················· | ···················· | ···················· | ···················· | |
| G | 4424 hCoV-19/Australia/VIC2708/2020\|EPI_ISL_521546\|2020 | ···················· | ···················· | ····················· | ···················· | ···················· | ···················· | |
| G | 4425 hCoV-19/Australia/VIC2710/2020\|EPI_ISL_521548\|2020 | ···················· | ···················· | ····················· | ···················· | ···················· | ···················· | |
| G | 4426 hCoV-19/Australia/VIC2711/2020\|EPI_ISL_521549\|2020 | ···················· | ···················· | ····················· | ···················· | ···················· | ···················· | |
| G | 4427 hCoV-19/Australia/VIC2712/2020\|EPI_ISL_521550\|2020 | ···················· | ···················· | ····················· | ···················· | ···················· | ···················· | |
| G | 4428 hCoV-19/Australia/VIC2713/2020\|EPI_ISL_521551\|2020 | ···················· | ···················· | ····················· | ···················· | ···················· | ···················· | |
| G | 4429 hCoV-19/Australia/VIC2714/2020\|EPI_ISL_521552\|2020 | ···················· | ···················· | ····················· | ···················· | ···················· | ···················· | |
| G | 4430 hCoV-19/Australia/VIC2716/2020\|EPI_ISL_521553\|2020 | ···················· | ···················· | ····················· | ···················· | ···················· | ···················· | |
| G | 4431 hCoV-19/Australia/VIC2717/2020\|EPI_ISL_521554\|2020 | ···················· | ···················· | ····················· | ···················· | ···················· | ···················· | |
| G | 4432 hCoV-19/Australia/VIC2718/2020\|EPI_ISL_521555\|2020 | ···················· | ···················· | ····················· | ···················· | ···················· | ···················· | |
| G | 4433 hCoV-19/Australia/VIC2719/2020\|EPI_ISL_521556\|2020 | ···················· | ···················· | ····················· | ···················· | ···················· | ···················· | |
| G | 4434 hCoV-19/Australia/VIC2720/2020\|EPI_ISL_521557\|2020 | ···················· | ···················· | ····················· | ···················· | ···················· | ···················· | |
| G | 4435 hCoV-19/Australia/VIC2721/2020\|EPI_ISL_521558\|2020 | ···················· | ···················· | ····················· | ···················· | ···················· | ···················· | |
| G | 4436 hCoV-19/Italy/VEN-IZSVe-60-40/2020\|EPI_ISL_522865\| | ···················· | ···················· | ····················· | ···················· | ···················· | ···················· | |
| G | 4437 hCoV-19/Italy/VEN-IZSVe-63-7/2020\|EPI_ISL_522866\|2 | ···················· | ···················· | ····················· | ···················· | ···················· | ···················· | |
| G | 4438 hCoV-19/Italy/VEN-IZSVe-71-10/2020\|EPI_ISL_522867\| | ···················· | ···················· | ····················· | ···················· | ···················· | ···················· | |
| G | 4439 hCoV-19/Italy/VEN-IZSVe-94-28/2020\|EPI_ISL_522868\| | ···················· | ···················· | ····················· | ···················· | ···················· | ···················· | |
| G | 4440 hCoV-19/USA/MD-MDH-0122/2020\|EPI_ISL_522869\|2020-0 | ·····G·············· | ···················· | ····················· | ···················· | ···················· | ···················· | |
| G | 4441 hCoV-19/USA/MD-MDH-0123/2020\|EPI_ISL_522870\|2020-0 | ···················· | ···················· | ····················· | ···················· | ···················· | ···················· | |
| G | 4442 hCoV-19/USA/MD-MDH-0124/2020\|EPI_ISL_522871\|2020-0 | ···················· | ···················· | ····················· | ···················· | ···················· | ···················· | |
| G | 4447 hCoV-19/USA/IL-UW-1147/2020\|EPI_ISL_522881\|2020-08 | ···················· | ·············A······ | ····················· | ···················· | ···················· | ···················· | |
| G | 4448 hCoV-19/USA/MN-UW-1130/2020\|EPI_ISL_522882\|2020-08 | ···················· | ···················· | ····················· | ···················· | ···················· | ···················· | |
| G | 4449 hCoV-19/USA/WI-UW-1125/2020\|EPI_ISL_522883\|2020-08 | ···················· | ···················· | ····················· | ···················· | ···················· | ···················· | |
| G | 4451 hCoV-19/USA/WI-UW-1127/2020\|EPI_ISL_522885\|2020-08 | ···················· | ···················· | ····················· | ···················· | ···················· | ···················· | |
| G | 4452 hCoV-19/USA/WI-UW-1128/2020\|EPI_ISL_522886\|2020-08 | ···················· | ···················· | ····················· | ···················· | ···················· | ···················· | |
| G | 4453 hCoV-19/USA/WI-UW-1129/2020\|EPI_ISL_522887\|2020-08 | ···················· | ···················· | ····················· | ···················· | ···················· | ···················· | |
| G | 4457 hCoV-19/USA/UT-UPHL-2009101/2020\|EPI_ISL_525102\|20 | ···················· | ···················· | ····················· | ···················· | ···················· | ···················· | |
| G | 4458 hCoV-19/USA/UT-UPHL-2009102/2020\|EPI_ISL_525103\|20 | ···················· | ···················· | ····················· | ···················· | ···················· | ···················· | |
| G | 4459 hCoV-19/USA/UT-UPHL-2009103/2020\|EPI_ISL_525104\|20 | ···················· | ···················· | ····················· | ···················· | ···················· | ···················· | |
| G | 4460 hCoV-19/USA/UT-UPHL-2009105/2020\|EPI_ISL_525106\|20 | ···················· | ···················· | ····················· | ···················· | ·········A·········· | ···················· | |
| G | 4461 hCoV-19/USA/UT-UPHL-2009108/2020\|EPI_ISL_525108\|20 | ···················· | ···················· | ····················· | ···················· | ···················· | ···················· | |
| G | 4462 hCoV-19/USA/UT-UPHL-2009109/2020\|EPI_ISL_525109\|20 | ···················· | ···················· | ····················· | ···················· | ···················· | ···················· | |
| G | 4463 hCoV-19/USA/UT-UPHL-2009110/2020\|EPI_ISL_525110\|20 | ···················· | ···················· | ····················· | ···················· | ···················· | ···················· | |
| G | 4464 hCoV-19/USA/UT-UPHL-2009129/2020\|EPI_ISL_525124\|20 | ···················· | ···················· | ····················· | ···················· | ···················· | ···················· | |
| G | 4465 hCoV-19/USA/UT-UPHL-2009133/2020\|EPI_ISL_525128\|20 | ···················· | ···················· | ····················· | ···················· | ···················· | ···················· | |
| G | 4466 hCoV-19/USA/UT-UPHL-2009134/2020\|EPI_ISL_525129\|20 | ···················· | ···················· | ····················· | ···················· | ···················· | ···················· | |
| G | 4467 hCoV-19/USA/UT-UPHL-2009136/2020\|EPI_ISL_525131\|20 | ···················· | ···················· | ····················· | ···················· | ···················· | ···················· | |
| G | 4468 hCoV-19/USA/UT-UPHL-2009139/2020\|EPI_ISL_525133\|20 | ···················· | ···················· | ····················· | ···················· | ···················· | ···················· | |
| G | 4469 hCoV-19/USA/UT-UPHL-2009143/2020\|EPI_ISL_525136\|20 | ···················· | ·A·················· | ····················· | ···················· | ···················· | ···················· | |
| G | 4470 hCoV-19/USA/UT-UPHL-2009148/2020\|EPI_ISL_525138\|20 | ···················· | ···················· | ····················· | ···················· | ···················· | ···················· | |
| G | 4471 hCoV-19/USA/UT-UPHL-2009153/2020\|EPI_ISL_525143\|20 | ···················· | ···················· | ····················· | ···················· | ···················· | ···················· | |
| G | 4472 hCoV-19/USA/UT-UPHL-2009159/2020\|EPI_ISL_525148\|20 | ···················· | ···················· | ····················· | ···················· | ···················· | ···················· | |
| G | 4473 hCoV-19/USA/UT-UPHL-2009160/2020\|EPI_ISL_525149\|20 | ···················· | ···················· | ····················· | ···················· | ···················· | ···················· | |
| G | 4474 hCoV-19/USA/UT-UPHL-2009162/2020\|EPI_ISL_525150\|20 | ···················· | ···················· | ····················· | ···················· | ···················· | ···················· | |
| G | 4475 hCoV-19/USA/UT-UPHL-2009163/2020\|EPI_ISL_525151\|20 | ···················· | ···················· | ····················· | ···················· | ···················· | ···················· | |

| | | | | | | | | |
|---|---|---|---|---|---|---|---|---|
| G | 4476 hCoV-19/USA/UT-UPHL-2009165/2020\|EPI_ISL_525152\|20 | .................... | .................... | ..................... | .................... | .................... | .................... | |
| G | 4477 hCoV-19/USA/UT-UPHL-2009168/2020\|EPI_ISL_525154\|20 | .................... | .................... | ..................... | .................... | .................... | .................... | |
| G | 4478 hCoV-19/USA/UT-UPHL-2009169/2020\|EPI_ISL_525155\|20 | .................... | .................... | ..................... | .................... | .................... | .................... | |
| G | 4479 hCoV-19/USA/UT-UPHL-2009172/2020\|EPI_ISL_525158\|20 | .................... | .................... | ..................... | .................... | .................... | .................... | |
| G | 4480 hCoV-19/USA/UT-UPHL-2009173/2020\|EPI_ISL_525159\|20 | .................... | .................... | ..................... | .................... | .................... | .................... | |
| G | 4481 hCoV-19/USA/UT-UPHL-2009174/2020\|EPI_ISL_525160\|20 | .................... | .................... | ..................... | .................... | .................... | .................... | |
| G | 4482 hCoV-19/USA/UT-UPHL-2009176/2020\|EPI_ISL_525162\|20 | .................... | .................... | ..................... | .................... | .................... | .................... | |
| G | 4483 hCoV-19/USA/UT-UPHL-2009183/2020\|EPI_ISL_525166\|20 | .................... | .................... | ..................... | .................... | .................... | .................... | |
| G | 4484 hCoV-19/USA/UT-UPHL-2009190/2020\|EPI_ISL_525173\|20 | .................... | .................... | ..................... | .................... | .................... | .................... | |
| G | 4485 hCoV-19/USA/UT-UPHL-2009194/2020\|EPI_ISL_525177\|20 | .................... | .................... | ..................... | .................... | .................... | .................... | |
| G | 4486 hCoV-19/England/MILK-977B64/2020\|EPI_ISL_526401\|20 | .................... | .................... | ..................... | .................... | .................... | .................... | |
| G | 4487 hCoV-19/England/MILK-977DAA/2020\|EPI_ISL_526402\|20 | .................... | .................... | ..................... | .................... | .................... | .................... | |
| G | 4488 hCoV-19/England/MILK-977DC8/2020\|EPI_ISL_526403\|20 | .................... | .................... | ..................... | .................... | .................... | .................... | |
| G | 4489 hCoV-19/England/MILK-9781E9/2020\|EPI_ISL_526404\|20 | .................... | .A.................. | ..................... | .................... | .................... | .................... | |
| G | 4490 hCoV-19/England/MILK-97828C/2020\|EPI_ISL_526405\|20 | .................... | .................... | ..................... | .................... | .................... | .................... | |
| G | 4491 hCoV-19/England/MILK-9782D7/2020\|EPI_ISL_526406\|20 | .................... | .................... | ..................... | .................... | .................... | .................... | |
| G | 4492 hCoV-19/England/MILK-9782E6/2020\|EPI_ISL_526407\|20 | .................... | .................... | ..................... | .................... | .................... | .................... | |
| G | 4493 hCoV-19/England/MILK-9782F5/2020\|EPI_ISL_526408\|20 | .................... | .................... | ..................... | .................... | .................... | .................... | |
| G | 4494 hCoV-19/England/MILK-97832F/2020\|EPI_ISL_526410\|20 | .................... | .................... | ..................... | .................... | .................... | .................... | |
| G | 4495 hCoV-19/England/MILK-97833E/2020\|EPI_ISL_526411\|20 | .................... | .................... | ..................... | .................... | .................... | .................... | |
| G | 4496 hCoV-19/England/MILK-978389/2020\|EPI_ISL_526412\|20 | .................... | .................... | ..................... | .................... | .................... | .................... | |
| G | 4497 hCoV-19/England/MILK-978477/2020\|EPI_ISL_526413\|20 | .................... | .................... | ..................... | .................... | .................... | .................... | |
| G | 4498 hCoV-19/England/MILK-978486/2020\|EPI_ISL_526414\|20 | .................... | .................... | ..................... | .................... | .................... | .................... | |
| G | 4499 hCoV-19/England/MILK-978495/2020\|EPI_ISL_526415\|20 | .................... | .................... | ..................... | .................... | .................... | .................... | |
| G | 4500 hCoV-19/England/MILK-978583/2020\|EPI_ISL_526418\|20 | .................... | .................... | ..................... | .................... | .................... | .................... | |
| G | 4501 hCoV-19/England/MILK-9785A1/2020\|EPI_ISL_526419\|20 | .................... | .................... | ..................... | .................... | .................... | .................... | |
| G | 4502 hCoV-19/England/MILK-9785B0/2020\|EPI_ISL_526420\|20 | .................... | .................... | ..................... | .................... | .................... | .................... | |
| G | 4503 hCoV-19/England/MILK-9785CF/2020\|EPI_ISL_526421\|20 | .................... | .................... | ..................... | .................... | .................... | .................... | |
| G | 4504 hCoV-19/England/MILK-9785DE/2020\|EPI_ISL_526422\|20 | .................... | .................... | ..................... | .................... | .................... | .................... | |
| G | 4505 hCoV-19/England/MILK-978AB1/2020\|EPI_ISL_526423\|20 | .................... | .................... | ..................... | .................... | .................... | .................... | |
| G | 4506 hCoV-19/England/MILK-978AC0/2020\|EPI_ISL_526424\|20 | .................... | .................... | ..................... | .................... | .................... | .................... | |
| G | 4507 hCoV-19/England/NOTT-112F1C/2020\|EPI_ISL_526425\|20 | .................... | .................... | ..................... | .................... | .................... | .................... | |
| G | 4508 hCoV-19/England/NOTT-112F3A/2020\|EPI_ISL_526427\|20 | .................... | .................... | ..................... | .................... | .................... | .................... | |
| G | 4509 hCoV-19/England/NOTT-112F67/2020\|EPI_ISL_526430\|20 | .................... | .................... | ..................... | .................... | .................... | .................... | |
| G | 4510 hCoV-19/England/PORT-2D13C1/2020\|EPI_ISL_526434\|20 | .................... | .................... | ..................... | .................... | .................... | .................... | |
| G | 4511 hCoV-19/England/PORT-2D1FBE/2020\|EPI_ISL_526435\|20 | .................... | .................... | ..................... | .................... | .................... | .................... | |
| G | 4513 hCoV-19/Scotland/CVR3969/2020\|EPI_ISL_526458\|2020- | .................... | .................... | ..................... | .................... | .................... | .................... | |
| G | 4514 hCoV-19/Scotland/EDB1491/2020\|EPI_ISL_526459\|2020- | .................... | .A.................. | ..................... | .................... | .................... | .................... | |
| G | 4516 hCoV-19/Scotland/EDB1729/2020\|EPI_ISL_526465\|2020- | .................... | .A.................. | ..................... | .................... | .................... | .................... | |
| G | 4517 hCoV-19/Scotland/EDB1757/2020\|EPI_ISL_526466\|2020- | .................... | .A.................. | ..................... | .................... | .................... | .................... | |
| G | 4519 hCoV-19/Scotland/EDB1785/2020\|EPI_ISL_526469\|2020- | .................... | .A.................. | ..................... | .................... | .................... | .................... | |
| G | 4520 hCoV-19/Scotland/EDB2889/2020\|EPI_ISL_526470\|2020- | .................... | .................... | ..................... | .................... | .................... | .................... | |
| G | 4521 hCoV-19/Scotland/EDB3080/2020\|EPI_ISL_526471\|2020- | .................... | ...................A | ..................... | .................... | .................... | .................... | 1 |
| G | 4522 hCoV-19/Scotland/EDB3584/2020\|EPI_ISL_526472\|2020- | .................... | .................... | ..................... | .................... | .................... | .................... | |
| G | 4523 hCoV-19/Scotland/EDB3785/2020\|EPI_ISL_526473\|2020- | .................... | .A.................. | ..................... | .................... | .................... | .................... | |
| G | 4524 hCoV-19/Scotland/EDB3829/2020\|EPI_ISL_526474\|2020- | .................... | .................... | ..................... | .................... | .................... | .................... | |
| G | 4525 hCoV-19/Scotland/EDB4001/2020\|EPI_ISL_526475\|2020- | .................... | .................... | ..................... | .................... | .................... | .................... | |
| G | 4526 hCoV-19/Scotland/EDB4060/2020\|EPI_ISL_526476\|2020- | .................... | ...................A | ..................... | .................... | .................... | .................... | 1 |
| G | 4527 hCoV-19/Scotland/EDB4090/2020\|EPI_ISL_526477\|2020- | .................... | .................... | ..................... | .................... | .................... | .................... | |
| G | 4528 hCoV-19/Scotland/EDB4133/2020\|EPI_ISL_526478\|2020- | .................... | .................... | ..................... | .................... | .................... | .................... | |
| G | 4529 hCoV-19/Scotland/EDB4247/2020\|EPI_ISL_526479\|2020- | .................... | ...................A | ..................... | .................... | .................... | .................... | 1 |
| G | 4530 hCoV-19/Scotland/EDB4386/2020\|EPI_ISL_526480\|2020- | .................... | .................... | ..................... | .................... | .................... | .................... | |
| G | 4531 hCoV-19/Scotland/EDB4387/2020\|EPI_ISL_526481\|2020- | .................... | .................... | ..................... | .................... | .................... | .................... | |
| G | 4532 hCoV-19/Scotland/EDB4388/2020\|EPI_ISL_526482\|2020- | .................... | .................... | ..................... | .................... | .................... | .................... | |
| G | 4533 hCoV-19/Scotland/EDB4389/2020\|EPI_ISL_526483\|2020- | .................... | .................... | ..................... | .................... | .................... | .................... | |
| G | 4534 hCoV-19/Scotland/EDB4568/2020\|EPI_ISL_526484\|2020- | .................... | .................... | ..................... | .................... | .................... | .................... | |
| G | 4535 hCoV-19/Scotland/EDB5837/2020\|EPI_ISL_526485\|2020- | .................... | .................... | ..................... | .................... | .................... | .................... | |
| G | 4536 hCoV-19/Scotland/EDB5905/2020\|EPI_ISL_526486\|2020- | .................... | .................... | ..................... | .................... | .................... | .................... | |
| G | 4537 hCoV-19/Scotland/EDB5973/2020\|EPI_ISL_526487\|2020- | .................... | .................... | ..................... | .................... | .................... | .................... | |
| G | 4538 hCoV-19/Scotland/EDB5985/2020\|EPI_ISL_526488\|2020- | .................... | .................... | ..................... | .................... | .................... | .................... | |
| G | 4539 hCoV-19/Scotland/EDB5987/2020\|EPI_ISL_526489\|2020- | .................... | .................... | ..................... | .................... | .................... | .................... | |
| G | 4540 hCoV-19/Scotland/EDB6859/2020\|EPI_ISL_526494\|2020- | .................... | .................... | ..................... | .................... | .................... | .................... | |
| G | 4541 hCoV-19/Scotland/EDB6880/2020\|EPI_ISL_526495\|2020- | .................... | .A.................. | ..................... | .................... | .................... | .................... | |
| G | 4542 hCoV-19/Scotland/EDB6881/2020\|EPI_ISL_526496\|2020- | .................... | .A.................. | ..................... | .................... | .................... | .................... | |
| G | 4543 hCoV-19/Scotland/EDB6882/2020\|EPI_ISL_526497\|2020- | .................... | .A.................. | ..................... | .................... | .................... | .................... | |
| G | 4544 hCoV-19/Scotland/EDB6883/2020\|EPI_ISL_526498\|2020- | .................... | .A.................. | ..................... | .................... | .................... | .................... | |
| G | 4545 hCoV-19/Scotland/EDB6884/2020\|EPI_ISL_526499\|2020- | .................... | .A.................. | ..................... | .................... | .................... | .................... | |
| G | 4546 hCoV-19/USA/MN-CDC-6315/2020\|EPI_ISL_527700\|2020-0 | .................... | .A.................. | ..................... | .................... | .................... | .................... | |
| G | 4547 hCoV-19/USA/MN-CDC-6318/2020\|EPI_ISL_527701\|2020-0 | .................... | .................... | ..................... | .................... | .................... | .................... | |
| G | 4548 hCoV-19/USA/MN-CDC-6326/2020\|EPI_ISL_527702\|2020-0 | .................... | .................... | ..................... | .................... | .................... | .................... | |
| G | 4549 hCoV-19/USA/MN-CDC-6332/2020\|EPI_ISL_527703\|2020-0 | .................... | .................... | ..................... | .................... | .................... | .................... | |
| G | 4550 hCoV-19/USA/MN-CDC-6335/2020\|EPI_ISL_527704\|2020-0 | .................... | .................... | ..................... | .................... | .................... | .................... | |
| G | 4551 hCoV-19/USA/MN-CDC-6338/2020\|EPI_ISL_527705\|2020-0 | .................... | .................... | ..................... | .................... | .................... | .................... | |
| G | 4552 hCoV-19/USA/MN-CDC-6348/2020\|EPI_ISL_527706\|2020-0 | .................... | .................... | ..................... | .................... | .................... | .................... | |
| G | 4553 hCoV-19/USA/MN-CDC-6354/2020\|EPI_ISL_527707\|2020-0 | .................... | .................... | ..................... | .................... | .................... | .................... | |
| G | 4554 hCoV-19/USA/MN-CDC-6361/2020\|EPI_ISL_527708\|2020-0 | .................... | .................... | ..................... | .................... | .................... | .................... | |
| G | 4555 hCoV-19/USA/MN-CDC-6364/2020\|EPI_ISL_527709\|2020-0 | .................... | .................... | ..................... | .................... | .................... | .................... | |
| G | 4556 hCoV-19/USA/MN-CDC-6495/2020\|EPI_ISL_527710\|2020-0 | .................... | .................... | ..................... | .................... | .................... | .................... | |
| G | 4557 hCoV-19/USA/MN-CDC-6498/2020\|EPI_ISL_527711\|2020-0 | .................... | .................... | ..................... | .................... | .................... | .................... | |
| G | 4558 hCoV-19/USA/MN-CDC-6501/2020\|EPI_ISL_527712\|2020-0 | .................... | .................... | ..................... | .................... | .................... | .................... | |

```
G    4559 hCoV-19/USA/MN-CDC-6502/2020|EPI_ISL_527713|2020-0    ....................  ....................  .....................  ....................  ....................  ....................
G    4560 hCoV-19/USA/MN-CDC-6508/2020|EPI_ISL_527714|2020-0    ....................  ....................  .....................  ....................  ....................  ....................
G    4561 hCoV-19/USA/MN-CDC-6510/2020|EPI_ISL_527715|2020-0    ....................  ....................  .....................  ....................  ....................  ....................
G    4562 hCoV-19/USA/MN-CDC-6513/2020|EPI_ISL_527716|2020-0    ....................  ....................  .....................  ....................  ....................  ....................
G    4563 hCoV-19/USA/MN-CDC-6514/2020|EPI_ISL_527717|2020-0    ....................  ....................  .....................  ....................  ....................  ....................
G    4564 hCoV-19/USA/MN-CDC-6517/2020|EPI_ISL_527718|2020-0    ....................  ....................  .....................  ....................  ....................  ....................
G    4565 hCoV-19/USA/MN-CDC-6520/2020|EPI_ISL_527719|2020-0    ....................  ....................  .....................  ....................  ....................  ....................
G    4566 hCoV-19/USA/MN-CDC-6527/2020|EPI_ISL_527720|2020-0    ....................  ....................  .....................  ....................  ....................  ....................
G    4567 hCoV-19/USA/MN-CDC-6532/2020|EPI_ISL_527721|2020-0    ....................  ....................  .....................  ....................  ....................  ....................
G    4568 hCoV-19/USA/MN-CDC-6535/2020|EPI_ISL_527722|2020-0    ....................  ....................  .....................  ....................  ....................  ....................
G    4569 hCoV-19/USA/MN-CDC-6536/2020|EPI_ISL_527723|2020-0    ....................  ....................  .....................  ....................  ....................  ....................
G    4570 hCoV-19/USA/MN-CDC-6538/2020|EPI_ISL_527724|2020-0    ....................  ....................  .....................  ....................  ....................  ....................
G    4571 hCoV-19/USA/MN-CDC-6542/2020|EPI_ISL_527725|2020-0    ....................  ....................  ...A.................  ....................  ....................  ....................
G    4572 hCoV-19/USA/MN-CDC-6543/2020|EPI_ISL_527726|2020-0    ....................  ....................  ...A.................  ....................  ....................  ....................
G    4573 hCoV-19/USA/MN-CDC-6546/2020|EPI_ISL_527727|2020-0    ....................  ....................  .....................  ....................  ....................  ....................
G    4574 hCoV-19/USA/MN-CDC-6551/2020|EPI_ISL_527728|2020-0    ....................  ....................  .....................  ....................  ....................  ....................
G    4575 hCoV-19/USA/MN-CDC-6555/2020|EPI_ISL_527729|2020-0    ....................  ....................  .....................  ....................  ....................  ....................
G    4576 hCoV-19/USA/MN-CDC-6562/2020|EPI_ISL_527730|2020-0    ....................  ....................  .....................  ....................  ....................  ....................
G    4577 hCoV-19/USA/MN-CDC-6566/2020|EPI_ISL_527731|2020-0    ....................  ....................  .....................  ....................  ....................  ....................
G    4578 hCoV-19/USA/MN-CDC-6568/2020|EPI_ISL_527732|2020-0    ....................  ....................  .....................  ....................  ....................  ....................
G    4579 hCoV-19/USA/MN-CDC-6572/2020|EPI_ISL_527733|2020-0    ....................  ....................  .....................  ....................  ....................  ....................
G    4580 hCoV-19/USA/MN-CDC-6573/2020|EPI_ISL_527734|2020-0    ....................  ....................  .....................  ....................  ....................  ....................
G    4581 hCoV-19/USA/MN-CDC-6593/2020|EPI_ISL_527735|2020-0    ....................  ....................  .....................  ....................  ....................  ....................
G    4582 hCoV-19/USA/MN-CDC-6597/2020|EPI_ISL_527736|2020-0    ....................  ....................  .....................  ....................  ....................  ....................
G    4583 hCoV-19/USA/WI-CDC-0949/2020|EPI_ISL_527737|2020-0    ....................  ....................  .....................  ....................  ....................  ....................
G    4584 hCoV-19/Costa Rica/INC-0071/2020|EPI_ISL_527739|20    ....................  ....................  .....................  ....................  ....................  ....................
G    4585 hCoV-19/Costa Rica/INC-0072/2020|EPI_ISL_527740|20    ....................  ....................  .....................  ....................  ....................  ....................
G    4586 hCoV-19/Costa Rica/INC-0073/2020|EPI_ISL_527741|20    ....................  ....................  .....................  ....................  ....................  ....................
G    4587 hCoV-19/Costa Rica/INC-0074/2020|EPI_ISL_527742|20    ....................  ....................  .....................  ....................  ....................  ....................
G    4588 hCoV-19/Costa Rica/INC-0075/2020|EPI_ISL_527743|20    ....................  ....................  .....................  ....................  ....................  ....................
G    4589 hCoV-19/Costa Rica/INC-0076/2020|EPI_ISL_527744|20    ....................  ....................  .....................  ....................  ....................  ....................
G    4590 hCoV-19/Costa Rica/INC-0078/2020|EPI_ISL_527746|20    ....................  ....................  .....................  ....................  ....................  ....................
G    4591 hCoV-19/Costa Rica/INC-0079/2020|EPI_ISL_527747|20    ....................  ....................  .....................  ....................  ....................  ....................
G    4592 hCoV-19/Costa Rica/INC-0080/2020|EPI_ISL_527748|20    ....................  ....................  .....................  ....................  ....................  ....................
G    4593 hCoV-19/Costa Rica/INC-0081/2020|EPI_ISL_527749|20    ....................  ....................  .....................  ....................  ....................  ....................
G    4594 hCoV-19/Costa Rica/INC-0082/2020|EPI_ISL_527750|20    ....................  ....................  .....................  ....................  ....................  ....................
G    4595 hCoV-19/Costa Rica/INC-0083/2020|EPI_ISL_527751|20    ....................  ....................  .....................  ....................  ....................  ....................
G    4596 hCoV-19/Costa Rica/INC-0084/2020|EPI_ISL_527752|20    ....................  ....................  .....................  ....................  ....................  ....................
G    4597 hCoV-19/Costa Rica/INC-0085/2020|EPI_ISL_527753|20    ....................  ....................  .....................  ....................  ....................  ....................
G    4598 hCoV-19/Costa Rica/INC-0086/2020|EPI_ISL_527754|20    ....................  ....................  .....................  ....................  ....................  ....................
G    4599 hCoV-19/Costa Rica/INC-0087/2020|EPI_ISL_527755|20    ....................  ....................  .....................  ....................  ....................  ....................
G    4600 hCoV-19/Costa Rica/INC-0088/2020|EPI_ISL_527756|20    ....................  ....................  .....................  ....................  ....................  ....................
G    4601 hCoV-19/Costa Rica/INC-0089/2020|EPI_ISL_527757|20    ....................  ....................  .....................  ....................  ....................  ....................
G    4602 hCoV-19/Peru/ARE-INS-075/2020|EPI_ISL_527787|2020-    ....................  ....................  .....................  ....................  ....................  ....................
G    4603 hCoV-19/Peru/ARE-INS-076/2020|EPI_ISL_527789|2020-    ....................  ....................  .....................  ....................  ....................  ....................
G    4609 hCoV-19/Singapore/837/2020|EPI_ISL_518001|2020-07-    ....................  ....................  .....................  ....................  ....................  ....................
G    4610 hCoV-19/Singapore/838/2020|EPI_ISL_518002|2020-07-    ....................  ....................  .....................  ....................  ....................  ....................
G    4611 hCoV-19/Singapore/839/2020|EPI_ISL_518003|2020-07-    ....................  ....................  .....................  ....................  ....................  ....................
G    4612 hCoV-19/Singapore/840/2020|EPI_ISL_518004|2020-07-    ....................  ....................  .....................  ....................  ....................  ....................
G    4613 hCoV-19/Singapore/841/2020|EPI_ISL_518005|2020-07-    ....................  ....................  .....................  ....................  ....................  ....................
G    4614 hCoV-19/Singapore/842/2020|EPI_ISL_518006|2020-07-    ....................  ....................  .....................  ....................  ....................  ....................
G    4615 hCoV-19/Singapore/843/2020|EPI_ISL_518007|2020-07-    ....................  ....................  .....................  ....................  ....................  ....................
G    4616 hCoV-19/Singapore/844/2020|EPI_ISL_518008|2020-07-    ....................  ....................  .....................  ....................  ....................  ....................
G    4617 hCoV-19/Singapore/845/2020|EPI_ISL_518009|2020-07-    ....................  ....................  .....................  ....................  ....................  ....................
G    4618 hCoV-19/Australia/VIC4926/2020|EPI_ISL_519300|2020    ....................  ....................  .....................  ....................  ....................  ....................
G    4619 hCoV-19/Australia/VIC4928/2020|EPI_ISL_519302|2020    ....................  ....................  .....................  ....................  ....................  ....................
G    4620 hCoV-19/Australia/VIC4929/2020|EPI_ISL_519303|2020    ....................  ....................  .....................  ....................  ....................  ....................
G    4621 hCoV-19/Australia/VIC4930/2020|EPI_ISL_519304|2020    ....................  ....................  .....................  ....................  ....................  ....................
G    4622 hCoV-19/Australia/VIC4931/2020|EPI_ISL_519305|2020    ....................  ....................  .....................  ....................  ....................  ....................
G    4623 hCoV-19/Australia/VIC4932/2020|EPI_ISL_519306|2020    ....................  ....................  .....................  ....................  ....................  ....................
G    4624 hCoV-19/Australia/VIC4933/2020|EPI_ISL_519307|2020    ....................  ....................  .....................  ....................  ....................  ....................
G    4625 hCoV-19/Australia/VIC4934/2020|EPI_ISL_519308|2020    ....................  ....................  .....................  ....................  ....................  ....................
G    4626 hCoV-19/Australia/VIC4935/2020|EPI_ISL_519309|2020    ....................  ....................  .....................  ....................  ....................  ....................
G    4627 hCoV-19/Australia/VIC4936/2020|EPI_ISL_519310|2020    ....................  ....................  .....................  ....................  ....................  ....................
G    4628 hCoV-19/Australia/VIC4937/2020|EPI_ISL_519311|2020    ....................  ....................  .....................  ....................  ....................  ....................
G    4629 hCoV-19/Australia/VIC4938/2020|EPI_ISL_519312|2020    ....................  ....................  .....................  ....................  ....................  ....................
G    4630 hCoV-19/Australia/VIC4939/2020|EPI_ISL_519313|2020    ....................  ....................  .....................  ....................  ....................  ....................
G    4631 hCoV-19/Australia/VIC4940/2020|EPI_ISL_519314|2020    ....................  ....................  .....................  ....................  ....................  ....................
G    4632 hCoV-19/Australia/VIC4941/2020|EPI_ISL_519315|2020    ....................  ....................  .....................  ....................  ....................  ....................
G    4633 hCoV-19/Australia/VIC4943/2020|EPI_ISL_519317|2020    ....................  ....................  .....................  ....................  ....................  ....................
G    4634 hCoV-19/Australia/VIC4945/2020|EPI_ISL_519318|2020    ....................  ....................  .....................  ....................  ....................  ....................
G    4635 hCoV-19/Australia/VIC4946/2020|EPI_ISL_519319|2020    ....................  ....................  .....................  ....................  ....................  ....................
G    4636 hCoV-19/Australia/VIC4947/2020|EPI_ISL_519320|2020    ....................  ....................  .....................  ....................  ....................  ....................
G    4637 hCoV-19/Australia/VIC4948/2020|EPI_ISL_519321|2020    ....................  ....................  .....................  ....................  ....................  ....................
G    4638 hCoV-19/Australia/VIC4949/2020|EPI_ISL_519322|2020    ....................  ....................  .....................  ....................  ....................  ....................
G    4639 hCoV-19/Australia/VIC4950/2020|EPI_ISL_519323|2020    ....................  ....................  .....................  ....................  ....................  ....................
G    4640 hCoV-19/Australia/VIC4953/2020|EPI_ISL_519324|2020    ....................  ....................  .....................  ....................  ....................  ....................
G    4641 hCoV-19/Australia/VIC4954/2020|EPI_ISL_519325|2020    ....................  ....................  .....................  ....................  ....................  ....................
G    4642 hCoV-19/Australia/VIC4958/2020|EPI_ISL_519326|2020    ....................  ....................  .....................  ....................  ....................  ....................
G    4643 hCoV-19/Australia/VIC4963/2020|EPI_ISL_519329|2020    ....................  ....................  .....................  ....................  ....................  ....................
```

| | | | | | | | | |
|---|---|---|---|---|---|---|---|---|
| G | 4644 hCoV-19/Australia/VIC4964/2020\|EPI_ISL_519330\|2020 | ···················· | ···················· | ···················· | ···················· | ···················· | ···················· | |
| G | 4645 hCoV-19/Australia/VIC4965/2020\|EPI_ISL_519331\|2020 | ···················· | ···················· | ···················· | ···················· | ···················· | ···················· | |
| G | 4646 hCoV-19/Australia/VIC4966/2020\|EPI_ISL_519332\|2020 | ···················· | ···················· | ···················· | ···················· | ···················· | ···················· | |
| G | 4647 hCoV-19/Australia/VIC4967/2020\|EPI_ISL_519333\|2020 | ···················· | ···················· | ···················· | ···················· | ···················· | ···················· | |
| G | 4648 hCoV-19/Australia/VIC4968/2020\|EPI_ISL_519334\|2020 | ···················· | ···················· | ···················· | ···················· | ···················· | ···················· | |
| G | 4649 hCoV-19/Australia/VIC4970/2020\|EPI_ISL_519336\|2020 | ···················· | ···················· | ···················· | ···················· | ···················· | ···················· | |
| G | 4650 hCoV-19/Australia/VIC4971/2020\|EPI_ISL_519337\|2020 | ···················· | ···················· | ···················· | ···················· | ···················· | ···················· | |
| G | 4651 hCoV-19/Australia/VIC4972/2020\|EPI_ISL_519338\|2020 | ···················· | ···················· | ···················· | ···················· | ···················· | ···················· | |
| G | 4652 hCoV-19/Australia/VIC4973/2020\|EPI_ISL_519339\|2020 | ···················· | ···················· | ···················· | ···················· | ···················· | ···················· | |
| G | 4653 hCoV-19/Australia/VIC3551/2020\|EPI_ISL_520300\|2020 | ···················· | ···················· | ···················· | ···················· | ···················· | ···················· | |
| G | 4654 hCoV-19/Australia/VIC3553/2020\|EPI_ISL_520302\|2020 | ···················· | ···················· | ···················· | ···················· | NNN················· | ···················· | 1 |
| G | 4655 hCoV-19/Australia/VIC3554/2020\|EPI_ISL_520303\|2020 | ···················· | ···················· | ···················· | ···················· | ···················· | ···················· | |
| G | 4656 hCoV-19/Australia/VIC3555/2020\|EPI_ISL_520304\|2020 | ···················· | ···················· | ···················· | ···················· | ···················· | ···················· | |
| G | 4657 hCoV-19/Australia/VIC3556/2020\|EPI_ISL_520305\|2020 | ···················· | ···················· | ···················· | ···················· | ···················· | ···················· | |
| G | 4658 hCoV-19/Australia/VIC3557/2020\|EPI_ISL_520306\|2020 | ···················· | ···················· | ···················· | ···················· | ···················· | ···················· | |
| G | 4659 hCoV-19/Australia/VIC3559/2020\|EPI_ISL_520308\|2020 | ···················· | ···················· | ···················· | ···················· | ···················· | ···················· | |
| G | 4660 hCoV-19/Australia/VIC3560/2020\|EPI_ISL_520309\|2020 | ···················· | ···················· | ···················· | ···················· | ···················· | ···················· | |
| G | 4661 hCoV-19/Australia/VIC3562/2020\|EPI_ISL_520311\|2020 | ···················· | ···················· | ···················· | ···················· | ···················· | ···················· | |
| G | 4662 hCoV-19/Australia/VIC3563/2020\|EPI_ISL_520312\|2020 | ···················· | ···················· | ···················· | ···················· | ···················· | ···················· | |
| G | 4663 hCoV-19/Australia/VIC3564/2020\|EPI_ISL_520313\|2020 | ···················· | ···················· | ···················· | ···················· | ···················· | ···················· | |
| G | 4664 hCoV-19/Australia/VIC3565/2020\|EPI_ISL_520314\|2020 | ···················· | ···················· | ···················· | ···················· | NNN················· | ···················· | 1 |
| G | 4665 hCoV-19/Australia/VIC3566/2020\|EPI_ISL_520315\|2020 | ···················· | ···················· | ···················· | ···················· | ···················· | ···················· | |
| G | 4666 hCoV-19/Australia/VIC3567/2020\|EPI_ISL_520316\|2020 | ···················· | ···················· | ···················· | ···················· | ···················· | ···················· | |
| G | 4667 hCoV-19/Australia/VIC3568/2020\|EPI_ISL_520317\|2020 | ···················· | ···················· | ···················· | ···················· | ···················· | ···················· | |
| G | 4668 hCoV-19/Australia/VIC3571/2020\|EPI_ISL_520320\|2020 | ···················· | ···················· | ···················· | ···················· | ···················· | ···················· | |
| G | 4669 hCoV-19/Australia/VIC3573/2020\|EPI_ISL_520322\|2020 | ···················· | ···················· | ···················· | ···················· | ···················· | ···················· | |
| G | 4671 hCoV-19/Australia/VIC2765/2020\|EPI_ISL_521600\|2020 | ···················· | ···················· | ···················· | ···················· | ···················· | ···················· | |
| G | 4673 hCoV-19/Australia/VIC2767/2020\|EPI_ISL_521602\|2020 | ···················· | ···················· | ···················· | ···················· | ···················· | ···················· | |
| G | 4674 hCoV-19/Australia/VIC2768/2020\|EPI_ISL_521603\|2020 | ···················· | ···················· | ···················· | ···················· | ···················· | ···················· | |
| G | 4675 hCoV-19/Australia/VIC2769/2020\|EPI_ISL_521604\|2020 | ···················· | ···················· | ···················· | ···················· | ···················· | ···················· | |
| G | 4676 hCoV-19/Australia/VIC2771/2020\|EPI_ISL_521606\|2020 | ···················· | ···················· | ···················· | ···················· | ···················· | ···················· | |
| G | 4677 hCoV-19/Australia/VIC2772/2020\|EPI_ISL_521607\|2020 | ···················· | ···················· | ···················· | ···················· | ···················· | ···················· | |
| G | 4678 hCoV-19/Australia/VIC2773/2020\|EPI_ISL_521608\|2020 | ···················· | ···················· | ···················· | ···················· | ···················· | ···················· | |
| G | 4679 hCoV-19/Australia/VIC2774/2020\|EPI_ISL_521609\|2020 | ···················· | ···················· | ···················· | ···················· | ···················· | ···················· | |
| G | 4681 hCoV-19/Australia/VIC2777/2020\|EPI_ISL_521611\|2020 | ···················· | ···················· | ···················· | ···················· | ···················· | ···················· | |
| G | 4682 hCoV-19/Australia/VIC2778/2020\|EPI_ISL_521612\|2020 | ···················· | ···················· | ···················· | ···················· | ···················· | ···················· | |
| G | 4683 hCoV-19/Australia/VIC2779/2020\|EPI_ISL_521613\|2020 | ···················· | ···················· | ···················· | ···················· | ···················· | ···················· | |
| G | 4685 hCoV-19/Australia/VIC2781/2020\|EPI_ISL_521615\|2020 | ···················· | ···················· | ···················· | ···················· | ···················· | ···················· | |
| G | 4688 hCoV-19/Australia/VIC2784/2020\|EPI_ISL_521618\|2020 | ···················· | ···················· | ···················· | ···················· | ···················· | ···················· | |
| G | 4690 hCoV-19/Australia/VIC2788/2020\|EPI_ISL_521622\|2020 | ···················· | ···················· | ···················· | ···················· | ···················· | ···················· | |
| G | 4691 hCoV-19/Australia/VIC2790/2020\|EPI_ISL_521624\|2020 | ···················· | ···················· | ···················· | ···················· | ···················· | ···················· | |
| G | 4693 hCoV-19/Australia/VIC2793/2020\|EPI_ISL_521626\|2020 | ···················· | ···················· | ···················· | ···················· | ···················· | ···················· | |
| G | 4697 hCoV-19/Australia/VIC2797/2020\|EPI_ISL_521630\|2020 | ···················· | ···················· | ···················· | ···················· | ···················· | ···················· | |
| G | 4698 hCoV-19/Australia/VIC2800/2020\|EPI_ISL_521631\|2020 | ···················· | ···················· | ···················· | ···················· | ···················· | ···················· | |
| G | 4699 hCoV-19/Australia/VIC2801/2020\|EPI_ISL_521632\|2020 | ···················· | ···················· | ···················· | ···················· | ···················· | ···················· | |
| G | 4702 hCoV-19/Australia/VIC2804/2020\|EPI_ISL_521635\|2020 | ···················· | ···················· | ···················· | ···················· | ···················· | ···················· | |
| G | 4703 hCoV-19/Australia/VIC2805/2020\|EPI_ISL_521636\|2020 | ···················· | ···················· | ···················· | ···················· | ···················· | ···················· | |
| G | 4704 hCoV-19/Australia/VIC2806/2020\|EPI_ISL_521637\|2020 | ···················· | ···················· | ···················· | ···················· | NNN················· | ···················· | 1 |
| G | 4705 hCoV-19/Australia/VIC2807/2020\|EPI_ISL_521638\|2020 | ···················· | ···················· | ···················· | ···················· | ···················· | ···················· | |
| G | 4706 hCoV-19/Australia/VIC2808/2020\|EPI_ISL_521639\|2020 | ···················· | ···················· | ···················· | ···················· | ···················· | ···················· | |
| G | 4707 hCoV-19/Australia/VIC2809/2020\|EPI_ISL_521640\|2020 | ···················· | ···················· | ···················· | ···················· | ···················· | ···················· | |
| G | 4708 hCoV-19/Australia/VIC2810/2020\|EPI_ISL_521641\|2020 | ···················· | ···················· | ···················· | ···················· | ···················· | ···················· | |
| G | 4710 hCoV-19/Australia/VIC2812/2020\|EPI_ISL_521643\|2020 | ···················· | ···················· | ···················· | ···················· | ···················· | ···················· | |
| G | 4711 hCoV-19/Australia/VIC2813/2020\|EPI_ISL_521644\|2020 | ···················· | ···················· | ···················· | ···················· | ···················· | ···················· | |
| G | 4712 hCoV-19/Australia/VIC2814/2020\|EPI_ISL_521645\|2020 | ···················· | ···················· | ···················· | ···················· | ···················· | ···················· | |
| G | 4713 hCoV-19/Australia/VIC2815/2020\|EPI_ISL_521646\|2020 | ···················· | ···················· | ···················· | ···················· | ···················· | ···················· | |
| G | 4714 hCoV-19/Australia/VIC2816/2020\|EPI_ISL_521647\|2020 | ···················· | ···················· | ···················· | ···················· | ···················· | ···················· | |
| G | 4715 hCoV-19/Australia/VIC2817/2020\|EPI_ISL_521648\|2020 | ···················· | ···················· | ···················· | ···················· | ···················· | ···················· | |
| G | 4717 hCoV-19/Australia/VIC2819/2020\|EPI_ISL_521650\|2020 | ···················· | ···················· | ···················· | ···················· | ···················· | ···················· | |
| G | 4718 hCoV-19/Australia/VIC2820/2020\|EPI_ISL_521651\|2020 | ···················· | ···················· | ···················· | ···················· | ···················· | ···················· | |
| G | 4719 hCoV-19/Australia/VIC2821/2020\|EPI_ISL_521652\|2020 | ···················· | ···················· | ···················· | ···················· | ···················· | ···················· | |
| G | 4720 hCoV-19/Australia/VIC2822/2020\|EPI_ISL_521653\|2020 | ···················· | ···················· | ···················· | ···················· | ···················· | ···················· | |
| G | 4721 hCoV-19/Australia/VIC2823/2020\|EPI_ISL_521654\|2020 | ···················· | ···················· | ···················· | ···················· | ···················· | ···················· | |
| G | 4725 hCoV-19/USA/WI-UW-1146/2020\|EPI_ISL_522903\|2020-08 | ···················· | ···················· | ···················· | ···················· | ···················· | ···················· | |
| G | 4731 hCoV-19/USA/WI-UW-1153/2020\|EPI_ISL_522909\|2020-08 | ···················· | ···················· | ···················· | ···················· | ···················· | ···················· | |
| G | 4735 hCoV-19/USA/WI-UW-1157/2020\|EPI_ISL_522913\|2020-08 | ···················· | ···················· | ···················· | ···················· | ···················· | ···················· | |
| G | 4736 hCoV-19/USA/WI-UW-1158/2020\|EPI_ISL_522914\|2020-08 | ···················· | ···················· | ···················· | ···················· | ···················· | ···················· | |
| G | 4743 hCoV-19/USA/WI-UW-1171/2020\|EPI_ISL_522927\|2020-08 | ···················· | ·A·················· | ···················· | ···················· | ···················· | ···················· | |
| G | 4745 hCoV-19/USA/WI-UW-1174/2020\|EPI_ISL_522929\|2020-08 | ···················· | ···················· | ···················· | ···················· | ···················· | ···················· | |
| G | 4747 hCoV-19/USA/WI-UW-1177/2020\|EPI_ISL_522932\|2020-08 | ···················· | ···················· | ···················· | ···················· | ···················· | ···················· | |
| G | 4748 hCoV-19/USA/WI-UW-1178/2020\|EPI_ISL_522933\|2020-08 | ···················· | ···················· | ···················· | ···················· | ···················· | ···················· | |
| G | 4749 hCoV-19/USA/WI-UW-1180/2020\|EPI_ISL_522935\|2020-08 | ···················· | ···················· | ···················· | ···················· | ···················· | ···················· | |
| G | 4750 hCoV-19/USA/WI-UW-1181/2020\|EPI_ISL_522936\|2020-08 | ···················· | ···················· | ···················· | ···················· | ···················· | ···················· | |
| G | 4751 hCoV-19/USA/WI-UW-1182/2020\|EPI_ISL_522937\|2020-08 | ···················· | ···················· | ···················· | ···················· | ···················· | ···················· | |
| G | 4752 hCoV-19/USA/WI-UW-1183/2020\|EPI_ISL_522938\|2020-07 | ···················· | ···················· | ···················· | ···················· | ···················· | ···················· | |
| G | 4753 hCoV-19/USA/WI-UW-1172/2020\|EPI_ISL_522939\|2020-08 | ···················· | ···················· | ···················· | ···················· | ···················· | ···················· | |
| G | 4757 hCoV-19/USA/TX-DSHS-0120/2020\|EPI_ISL_522949\|2020- | ···················· | ···················· | ···················· | ···················· | ···················· | ···················· | |
| G | 4758 hCoV-19/USA/TX-DSHS-0126/2020\|EPI_ISL_522953\|2020- | ···················· | ···················· | ···················· | ···················· | ···················· | ···················· | |
| G | 4759 hCoV-19/USA/TX-DSHS-0133/2020\|EPI_ISL_522960\|2020- | ···················· | ···················· | ···················· | ···················· | ···················· | ···················· | |
| G | 4760 hCoV-19/USA/TX-DSHS-0140/2020\|EPI_ISL_522966\|2020- | ···················· | ···················· | ···················· | ···················· | ···················· | ···················· | |

G 4761 hCoV-19/USA/TX-DSHS-0143/2020|EPI_ISL_522969|2020- ···················· ···················· ····················· ···················· ···················· ····················
G 4762 hCoV-19/USA/TX-DSHS-0145/2020|EPI_ISL_522971|2020- ···················· ···················· ····················· ···················· ···················· ····················
G 4767 hCoV-19/Scotland/GCVR-171B62/2020|EPI_ISL_524500|2 ···················· ···················· ····················· ···················· ···················· ····················
G 4768 hCoV-19/England/CAMB-7E498/2020|EPI_ISL_524501|202 ···················· ···················· ····················· ···················· ···················· ····················
G 4769 hCoV-19/England/CAMB-7E3AA/2020|EPI_ISL_524509|202 ···················· ···················· ····················· ···················· ···················· ····················
G 4770 hCoV-19/Scotland/GCVR-171E2D/2020|EPI_ISL_524513|2 ···················· ···················· ····················· ···················· ···················· ····················
G 4771 hCoV-19/Scotland/GCVR-173113/2020|EPI_ISL_524514|2 ···················· ············A······· ····················· ···················· ···················· ····················
G 4772 hCoV-19/Scotland/GCVR-172CB9/2020|EPI_ISL_524516|2 ···················· ···················A ····················· ···················· ···················· ···················· 1
G 4773 hCoV-19/Scotland/GCVR-170D5E/2020|EPI_ISL_524518|2 ···················· ···················· ····················· ···················· ···················· ····················
G 4774 hCoV-19/Scotland/GCVR-171582/2020|EPI_ISL_524519|2 ···················· ···················· ····················· ···················· ···················· ····················
G 4775 hCoV-19/Scotland/GCVR-171661/2020|EPI_ISL_524520|2 ···················· ·A·················· ····················· ···················· ···················· ····················
G 4776 hCoV-19/USA/UT-UPHL-2009197/2020|EPI_ISL_525180|20 ···················· ···················· ····················· ···················· ···················· ····················
G 4777 hCoV-19/USA/UT-UPHL-2009201/2020|EPI_ISL_525183|20 ···················· ···················· ····················· ···················· ···················· ····················
G 4778 hCoV-19/USA/UT-UPHL-2009204/2020|EPI_ISL_525186|20 ···················· ···················· ····················· ···················· ···················· ····················
G 4779 hCoV-19/USA/UT-UPHL-2009205/2020|EPI_ISL_525187|20 ···················· ·A·················· ····················· ···················· ···················· ····················
G 4780 hCoV-19/USA/VA-DCLS-1158/2020|EPI_ISL_525189|2020- ···················· ···················· ····················· ···················· ···················· ····················
G 4781 hCoV-19/USA/VA-DCLS-1159/2020|EPI_ISL_525190|2020- ···················· ···················· ····················· ···················· ···················· ····················
G 4782 hCoV-19/USA/VA-DCLS-1161/2020|EPI_ISL_525191|2020- ···················· ···················· ····················· ···················· ···················· ····················
G 4783 hCoV-19/USA/VA-DCLS-1164/2020|EPI_ISL_525192|2020- ···················· ···················· ····················· ···················· ···················· ····················
G 4784 hCoV-19/USA/VA-DCLS-1165/2020|EPI_ISL_525193|2020- ···················· ···················· ····················· ···················· ···················· ····················
G 4785 hCoV-19/USA/VA-DCLS-1167/2020|EPI_ISL_525194|2020- ···················· ···················· ····················· ···················· ···················· ····················
G 4786 hCoV-19/USA/VA-DCLS-1168/2020|EPI_ISL_525195|2020- ···················· ·A·················· ····················· ···················· ···················· ····················
G 4787 hCoV-19/USA/VA-DCLS-1169/2020|EPI_ISL_525196|2020- ···················· ·A·················· ····················· ···················· ···················· ····················
G 4788 hCoV-19/USA/VA-DCLS-1170/2020|EPI_ISL_525197|2020- ···················· ·A·················· ····················· ···················· ···················· ····················
G 4789 hCoV-19/USA/VA-DCLS-1171/2020|EPI_ISL_525198|2020- ···················· ·A·················· ····················· ···················· ···················· ····················
G 4790 hCoV-19/USA/VA-DCLS-1172/2020|EPI_ISL_525199|2020- ···················· ·A·················· ····················· ···················· ···················· ····················
G 4791 hCoV-19/USA/AK-OHSU-1669/2020|EPI_ISL_525802|2020- ···················· ···················· ····················· ···················· ···················· ····················
G 4792 hCoV-19/USA/CA-OHSU-0690/2020|EPI_ISL_525803|2020- ···················· ···················· ····················· ···················· ···················· ····················
G 4793 hCoV-19/USA/CA-OHSU-0857/2020|EPI_ISL_525804|2020- ···················· ·A·················· ····················· ···················· ···················· ····················
G 4794 hCoV-19/USA/CA-OHSU-0925/2020|EPI_ISL_525805|2020- ···················· ·A·················· ····················· ···················· ···················· ····················
G 4795 hCoV-19/USA/ID-OHSU-1977/2020|EPI_ISL_525806|2020- ···················· ···················· ····················· ···················· ···················· ····················
G 4796 hCoV-19/USA/OR-OHSU-0350/2020|EPI_ISL_525807|2020- ···················· ···················· ····················· ···················· ···················· ····················
G 4797 hCoV-19/USA/OR-OHSU-0440/2020|EPI_ISL_525808|2020- ···················· ···················· ····················· ···················· ···················· ····················
G 4798 hCoV-19/USA/OR-OHSU-0446/2020|EPI_ISL_525809|2020- ···················· ···················· ····················· ···················· ···················· ····················
G 4799 hCoV-19/USA/OR-OHSU-0456/2020|EPI_ISL_525813|2020- ···················· ···················· ····················· ···················· ···················· ····················
G 4800 hCoV-19/USA/OR-OHSU-0460/2020|EPI_ISL_525815|2020- ···················· ···················· ····················· ···················· ···················· ····················
G 4801 hCoV-19/USA/OR-OHSU-0466/2020|EPI_ISL_525816|2020- ···················· ···················· ····················· ···················· ···················· ····················
G 4802 hCoV-19/USA/OR-OHSU-0468/2020|EPI_ISL_525817|2020- ···················· ···················· ····················· ···················· ···················· ····················
G 4803 hCoV-19/USA/OR-OHSU-0470/2020|EPI_ISL_525818|2020- ···················· ···················· ····················· ···················· ···················· ····················
G 4804 hCoV-19/USA/OR-OHSU-0484/2020|EPI_ISL_525823|2020- ···················· ···················· ····················· ···················· ···················· ····················
G 4805 hCoV-19/USA/OR-OHSU-0503/2020|EPI_ISL_525825|2020- ···················· ···················· ····················· ···················· ···················· ····················
G 4806 hCoV-19/USA/OR-OHSU-0508/2020|EPI_ISL_525826|2020- ···················· ···················· ····················· ···················· ···················· ····················
G 4807 hCoV-19/USA/OR-OHSU-0509/2020|EPI_ISL_525827|2020- ···················· ···················· ····················· ···················· ···················· ····················
G 4808 hCoV-19/USA/OR-OHSU-0510/2020|EPI_ISL_525828|2020- ···················· ···················· ····················· ···················· ···················· ····················
G 4809 hCoV-19/USA/OR-OHSU-0514/2020|EPI_ISL_525829|2020- ···················· ···················· ····················· ···················· ···················· ····················
G 4810 hCoV-19/USA/OR-OHSU-0516/2020|EPI_ISL_525830|2020- ···················· ···················· ····················· ···················· ···················· ····················
G 4811 hCoV-19/USA/OR-OHSU-0526/2020|EPI_ISL_525831|2020- ···················· ···················· ····················· ···················· ···················· ····················
G 4812 hCoV-19/USA/OR-OHSU-0528/2020|EPI_ISL_525832|2020- ···················· ···················· ····················· ···················· ···················· ····················
G 4813 hCoV-19/USA/OR-OHSU-0529/2020|EPI_ISL_525833|2020- ···················· ···················· ····················· ···················· ···················· ····················
G 4814 hCoV-19/USA/OR-OHSU-0547/2020|EPI_ISL_525841|2020- ···················· ···················· ····················· ···················· ···················· ····················
G 4815 hCoV-19/USA/OR-OHSU-0548/2020|EPI_ISL_525842|2020- ···················· ···················· ····················· ···················· ···················· ····················
G 4816 hCoV-19/USA/OR-OHSU-0608/2020|EPI_ISL_525843|2020- ···················· ···················· ····················· ···················· ···················· ····················
G 4817 hCoV-19/USA/OR-OHSU-0612/2020|EPI_ISL_525845|2020- ···················· ···················· ····················· ···················· ···················· ····················
G 4818 hCoV-19/USA/OR-OHSU-0614/2020|EPI_ISL_525846|2020- ···················· ···················· ····················· ···················· ···················· ····················
G 4819 hCoV-19/USA/OR-OHSU-0615/2020|EPI_ISL_525847|2020- ···················· ···················· ····················· ···················· ···················· ····················
G 4820 hCoV-19/USA/OR-OHSU-0616/2020|EPI_ISL_525848|2020- ···················· ···················· ····················· ···················· ···················· ····················
G 4821 hCoV-19/USA/OR-OHSU-0617/2020|EPI_ISL_525849|2020- ···················· ···················· ····················· ···················· ···················· ····················
G 4822 hCoV-19/USA/OR-OHSU-0619/2020|EPI_ISL_525851|2020- ···················· ···················· ····················· ···················· ···················· ····················
G 4823 hCoV-19/USA/OR-OHSU-0620/2020|EPI_ISL_525852|2020- ···················· ···················· ····················· ···················· ···················· ····················
G 4824 hCoV-19/USA/OR-OHSU-0624/2020|EPI_ISL_525854|2020- ···················· ···················· ····················· ···················· ···················· ····················
G 4825 hCoV-19/USA/OR-OHSU-0627/2020|EPI_ISL_525856|2020- ···················· ···················· ····················· ···················· ···················· ····················
G 4826 hCoV-19/Switzerland/BL-UHB-42189907/2020|EPI_ISL_5 ···················· ···················· ····················· ···················· ···················· ····················
G 4827 hCoV-19/Switzerland/BS-UHB-42190027/2020|EPI_ISL_5 ···················· ···················· ····················· ···················· ···················· ····················
G 4828 hCoV-19/Switzerland/BS-UHB-42190051/2020|EPI_ISL_5 ···················· ·A·················· ····················· ···················· ···················· ····················
G 4829 hCoV-19/Switzerland/BS-UHB-42190083/2020|EPI_ISL_5 ···················· ···················· ····················· ···················· ···················· ····················
G 4830 hCoV-19/Switzerland/BS-UHB-42190125/2020|EPI_ISL_5 ···················· ···················· ····················· ···················· ···················· ····················
G 4831 hCoV-19/Switzerland/BS-UHB-42190173/2020|EPI_ISL_5 ···················· ···················· ····················· ···················· ···················· ····················
G 4832 hCoV-19/Switzerland/BS-UHB-42190251/2020|EPI_ISL_5 ···················· ···················· ····················· ···················· ···················· ····················
G 4833 hCoV-19/Switzerland/BL-UHB-42190253/2020|EPI_ISL_5 ···················· ···················· ····················· ···················· ···················· ····················
G 4834 hCoV-19/Switzerland/BL-UHB-42190269/2020|EPI_ISL_5 ···················· ···················· ····················· ···················· ···················· ····················
G 4835 hCoV-19/Switzerland/BS-UHB-42190282/2020|EPI_ISL_5 ···················· ···················· ····················· ···················· ···················· ····················
G 4836 hCoV-19/Switzerland/BS-UHB-42190292/2020|EPI_ISL_5 ···················· ···················· ····················· ···················· ···················· ····················
G 4837 hCoV-19/Switzerland/BS-UHB-42190293/2020|EPI_ISL_5 ···················· ···················· ····················· ···················· ···················· ····················
G 4838 hCoV-19/Switzerland/BS-UHB-42190359/2020|EPI_ISL_5 ···················· ···················· ····················· ···················· ···················· ····················
G 4839 hCoV-19/Switzerland/BS-UHB-42190378/2020|EPI_ISL_5 ···················· ···················· ····················· ···················· ···················· ····················
G 4840 hCoV-19/Switzerland/BL-UHB-42190565/2020|EPI_ISL_5 ···················· ···················· ····················· ···················· ···················· ····················
G 4841 hCoV-19/Switzerland/BL-UHB-42190581/2020|EPI_ISL_5 ···················· ···················· ····················· ···················· ···················· ····················
G 4842 hCoV-19/Switzerland/BS-UHB-42190622/2020|EPI_ISL_5 ···················· ···················· ····················· ···················· ···················· ····················
G 4843 hCoV-19/Switzerland/BS-UHB-42190675/2020|EPI_ISL_5 ···················· ···················· ····················· ···················· ···················· ····················
G 4844 hCoV-19/Switzerland/BS-UHB-42190783/2020|EPI_ISL_5 ···················· ···················· ····················· ···················· ···················· ····················

| | | | | | | | | |
|---|---|---|---|---|---|---|---|---|
| G | 4845 | hCoV-19/Switzerland/BL-UHB-42190803/2020\|EPI_ISL_5 | .................... | .................... | .................... | .................... | .................... | .................... |
| G | 4846 | hCoV-19/Switzerland/BL-UHB-42190921/2020\|EPI_ISL_5 | .................... | .................... | .................... | .................... | .................... | .................... |
| G | 4847 | hCoV-19/Switzerland/BS-UHB-42190958/2020\|EPI_ISL_5 | .................... | .................... | .................... | .................... | .................... | .................... |
| G | 4848 | hCoV-19/Switzerland/BS-UHB-42190971/2020\|EPI_ISL_5 | .................... | .................... | .................... | .................... | .................... | .................... |
| G | 4849 | hCoV-19/Switzerland/BS-UHB-42190980/2020\|EPI_ISL_5 | .................... | .................... | .................... | .................... | .................... | .................... |
| G | 4850 | hCoV-19/Switzerland/BS-UHB-42190994/2020\|EPI_ISL_5 | .................... | .................... | .................... | .................... | .................... | .................... |
| G | 4851 | hCoV-19/Switzerland/BL-UHB-42190998/2020\|EPI_ISL_5 | .................... | .................... | .................... | .................... | .................... | .................... |
| G | 4852 | hCoV-19/Switzerland/BS-UHB-42190999/2020\|EPI_ISL_5 | .................... | .................... | .................... | .................... | .................... | .................... |
| G | 4853 | hCoV-19/Switzerland/BL-UHB-42191004/2020\|EPI_ISL_5 | .................... | .................... | .................... | .................... | .................... | .................... |
| G | 4854 | hCoV-19/Switzerland/BS-UHB-42191012/2020\|EPI_ISL_5 | .................... | .................... | .................... | .................... | .................... | .................... |
| G | 4855 | hCoV-19/Switzerland/BL-UHB-42191063/2020\|EPI_ISL_5 | .................... | .................... | .................... | .................... | .................... | .................... |
| G | 4856 | hCoV-19/Switzerland/BS-UHB-42191134/2020\|EPI_ISL_5 | .................... | .................... | .................... | .................... | .................... | .................... |
| G | 4857 | hCoV-19/Switzerland/BL-UHB-42191153/2020\|EPI_ISL_5 | .................... | .................... | .................... | .................... | .................... | .................... |
| G | 4858 | hCoV-19/Switzerland/BS-UHB-42191158/2020\|EPI_ISL_5 | .................... | .................... | .................... | .................... | .................... | .................... |
| G | 4859 | hCoV-19/Switzerland/BS-UHB-42191222/2020\|EPI_ISL_5 | .................... | .................... | .................... | .................... | .................... | .................... |
| G | 4860 | hCoV-19/Switzerland/BS-UHB-42191225/2020\|EPI_ISL_5 | .................... | .................... | .................... | .................... | .................... | .................... |
| G | 4861 | hCoV-19/Switzerland/BL-UHB-42191355/2020\|EPI_ISL_5 | .................... | .................... | .................... | .................... | .................... | .................... |
| G | 4862 | hCoV-19/Switzerland/BS-UHB-42191430/2020\|EPI_ISL_5 | .................... | .................... | .................... | .................... | .................... | .................... |
| G | 4863 | hCoV-19/Switzerland/BS-UHB-42191432/2020\|EPI_ISL_5 | .................... | .................... | .................... | .................... | .................... | .................... |
| G | 4864 | hCoV-19/Switzerland/BL-UHB-42191497/2020\|EPI_ISL_5 | .................... | .................... | .................... | .................... | .................... | .................... |
| G | 4865 | hCoV-19/England/LIVE-AA16F/2020\|EPI_ISL_517410\|202 | .................... | .................... | .................... | .................... | .................... | .................... |
| G | 4866 | hCoV-19/England/LIVE-AA1C9/2020\|EPI_ISL_517416\|202 | .................... | .................... | .................... | .................... | .................... | .................... |
| G | 4870 | hCoV-19/England/LIVE-AA33C/2020\|EPI_ISL_517438\|202 | .................... | .................... | .................... | .................... | .................... | .................... |
| G | 4871 | hCoV-19/England/LIVE-AA6F7/2020\|EPI_ISL_517490\|202 | .................... | .................... | .................... | .................... | .................... | .................... |
| G | 4872 | hCoV-19/England/LIVE-AA703/2020\|EPI_ISL_517491\|202 | .................... | .................... | .................... | .................... | .................... | .................... |
| G | 4873 | hCoV-19/England/LIVE-AA75E/2020\|EPI_ISL_517496\|202 | .................... | .................... | .................... | .................... | .................... | .................... |
| G | 4874 | hCoV-19/England/LIVE-AA76D/2020\|EPI_ISL_517497\|202 | .................... | .................... | .................... | .................... | .................... | .................... |
| G | 4875 | hCoV-19/England/LIVE-AA77C/2020\|EPI_ISL_517498\|202 | .................... | .................... | .................... | .................... | .................... | .................... |
| G | 4876 | hCoV-19/Australia/VIC5921/2020\|EPI_ISL_518700\|2020 | .................... | .................... | .................... | .................... | .................... | .................... |
| G | 4877 | hCoV-19/Australia/VIC5922/2020\|EPI_ISL_518701\|2020 | .................... | .................... | .................... | .................... | .................... | .................... |
| G | 4878 | hCoV-19/Australia/VIC5923/2020\|EPI_ISL_518702\|2020 | .................... | .................... | .................... | .................... | .................... | .................... |
| G | 4879 | hCoV-19/Australia/VIC5924/2020\|EPI_ISL_518703\|2020 | .................... | .................... | .................... | .................... | .................... | .................... |
| G | 4880 | hCoV-19/Australia/VIC5925/2020\|EPI_ISL_518704\|2020 | .................... | .................... | .................... | .................... | .................... | .................... |
| G | 4881 | hCoV-19/Australia/VIC5926/2020\|EPI_ISL_518705\|2020 | .................... | .................... | .................... | .................... | .................... | .................... |
| G | 4882 | hCoV-19/Australia/VIC5927/2020\|EPI_ISL_518706\|2020 | .................... | .................... | .................... | .................... | .................... | .................... |
| G | 4883 | hCoV-19/Australia/VIC5928/2020\|EPI_ISL_518707\|2020 | .................... | .................... | .................... | .................... | .................... | .................... |
| G | 4884 | hCoV-19/Australia/VIC5929/2020\|EPI_ISL_518708\|2020 | .................... | .................... | .................... | .................... | .................... | .................... |
| G | 4885 | hCoV-19/Australia/VIC5930/2020\|EPI_ISL_518709\|2020 | .................... | .................... | .................... | .................... | .................... | .................... |
| G | 4886 | hCoV-19/Australia/VIC5932/2020\|EPI_ISL_518711\|2020 | .................... | .................... | .................... | .................... | .................... | .................... |
| G | 4887 | hCoV-19/Australia/VIC5933/2020\|EPI_ISL_518712\|2020 | .................... | .................... | .................... | .................... | .................... | .................... |
| G | 4888 | hCoV-19/Australia/VIC5934/2020\|EPI_ISL_518713\|2020 | .................... | .................... | .................... | .................... | .................... | .................... |
| G | 4890 | hCoV-19/Australia/VIC5936/2020\|EPI_ISL_518715\|2020 | .................... | .................... | .................... | .................... | .................... | .................... |
| G | 4891 | hCoV-19/Australia/VIC5937/2020\|EPI_ISL_518716\|2020 | .................... | .................... | .................... | .................... | .................... | .................... |
| G | 4892 | hCoV-19/Australia/VIC5938/2020\|EPI_ISL_518717\|2020 | .................... | .................... | .................... | .................... | .................... | .................... |
| G | 4893 | hCoV-19/Australia/VIC5939/2020\|EPI_ISL_518718\|2020 | .................... | .................... | .................... | .................... | .................... | .................... |
| G | 4894 | hCoV-19/Australia/VIC5940/2020\|EPI_ISL_518719\|2020 | .................... | .................... | .................... | .................... | .................... | .................... |
| G | 4895 | hCoV-19/Australia/VIC5941/2020\|EPI_ISL_518720\|2020 | .................... | .................... | .................... | .................... | .................... | .................... |
| G | 4896 | hCoV-19/Australia/VIC5942/2020\|EPI_ISL_518721\|2020 | .................... | .................... | .................... | .................... | .................... | .................... |
| G | 4897 | hCoV-19/Australia/VIC5943/2020\|EPI_ISL_518722\|2020 | .................... | .................... | .................... | .................... | .................... | .................... |
| G | 4898 | hCoV-19/Australia/VIC5944/2020\|EPI_ISL_518723\|2020 | .................... | .................... | .................... | .................... | .................... | .................... |
| G | 4899 | hCoV-19/Australia/VIC5945/2020\|EPI_ISL_518724\|2020 | .................... | .................... | .................... | .................... | .................... | .................... |
| G | 4900 | hCoV-19/Australia/VIC5947/2020\|EPI_ISL_518725\|2020 | .................... | .................... | .................... | .................... | .................... | .................... |
| G | 4901 | hCoV-19/Australia/VIC5950/2020\|EPI_ISL_518726\|2020 | .................... | .................... | .................... | .................... | .................... | .................... |
| G | 4902 | hCoV-19/Australia/VIC5951/2020\|EPI_ISL_518727\|2020 | .................... | .................... | .................... | .................... | .................... | .................... |
| G | 4903 | hCoV-19/Australia/VIC5952/2020\|EPI_ISL_518728\|2020 | .................... | .................... | .................... | .................... | .................... | .................... |
| G | 4904 | hCoV-19/Australia/VIC5953/2020\|EPI_ISL_518729\|2020 | .................... | .................... | .................... | .................... | .................... | .................... |
| G | 4905 | hCoV-19/Australia/VIC5954/2020\|EPI_ISL_518730\|2020 | .................... | .................... | .................... | .................... | .................... | .................... |
| G | 4906 | hCoV-19/Australia/VIC5955/2020\|EPI_ISL_518731\|2020 | .................... | .................... | .................... | .................... | .................... | .................... |
| G | 4907 | hCoV-19/Australia/VIC5957/2020\|EPI_ISL_518732\|2020 | .................... | .................... | .................... | .................... | .................... | .................... |
| G | 4908 | hCoV-19/Australia/VIC5958/2020\|EPI_ISL_518733\|2020 | .................... | .................... | .................... | .................... | .................... | .................... |
| G | 4909 | hCoV-19/Australia/VIC5959/2020\|EPI_ISL_518734\|2020 | .................... | .................... | .................... | .................... | .................... | .................... |
| G | 4910 | hCoV-19/Australia/VIC5960/2020\|EPI_ISL_518735\|2020 | .................... | .................... | .................... | .................... | .................... | .................... |
| G | 4911 | hCoV-19/Australia/VIC5961/2020\|EPI_ISL_518736\|2020 | .................... | .................... | .................... | .................... | .................... | .................... |
| G | 4912 | hCoV-19/Australia/VIC5962/2020\|EPI_ISL_518737\|2020 | .................... | .................... | .................... | .................... | .................... | .................... |
| G | 4913 | hCoV-19/Australia/VIC5963/2020\|EPI_ISL_518738\|2020 | .................... | .................... | .................... | .................... | .................... | .................... |
| G | 4914 | hCoV-19/Australia/VIC5964/2020\|EPI_ISL_518739\|2020 | .................... | .................... | .................... | .................... | .................... | .................... |
| G | 4915 | hCoV-19/Australia/VIC5965/2020\|EPI_ISL_518740\|2020 | .................... | .................... | .................... | .................... | .................... | .................... |
| G | 4916 | hCoV-19/Australia/VIC5966/2020\|EPI_ISL_518741\|2020 | .................... | .................... | .................... | .................... | .................... | .................... |
| G | 4917 | hCoV-19/Australia/VIC5967/2020\|EPI_ISL_518742\|2020 | .................... | .................... | .................... | .................... | .................... | .................... |
| G | 4918 | hCoV-19/Australia/VIC5968/2020\|EPI_ISL_518743\|2020 | .................... | .................... | .................... | .................... | .................... | .................... |
| G | 4919 | hCoV-19/Australia/VIC5969/2020\|EPI_ISL_518744\|2020 | .................... | .................... | .................... | .................... | .................... | .................... |
| G | 4920 | hCoV-19/Australia/VIC5970/2020\|EPI_ISL_518745\|2020 | .................... | .................... | .................... | .................... | .................... | .................... |
| G | 4921 | hCoV-19/Australia/VIC5971/2020\|EPI_ISL_518746\|2020 | .................... | .................... | .................... | .................... | .................... | .................... |
| G | 4922 | hCoV-19/Australia/VIC5972/2020\|EPI_ISL_518747\|2020 | .................... | .................... | .................... | .................... | .................... | .................... |
| G | 4923 | hCoV-19/Australia/VIC5973/2020\|EPI_ISL_518748\|2020 | .................... | .................... | .................... | .................... | .................... | .................... |
| G | 4924 | hCoV-19/Australia/VIC5974/2020\|EPI_ISL_518749\|2020 | .................... | .................... | .................... | .................... | .................... | .................... |
| G | 4925 | hCoV-19/Australia/VIC5975/2020\|EPI_ISL_518750\|2020 | .................... | .................... | .................... | .................... | .................... | .................... |
| G | 4926 | hCoV-19/Australia/VIC5976/2020\|EPI_ISL_518751\|2020 | .................... | .................... | .................... | .................... | .................... | .................... |
| G | 4927 | hCoV-19/Australia/VIC5977/2020\|EPI_ISL_518752\|2020 | .................... | .................... | .................... | .................... | .................... | .................... |
| G | 4928 | hCoV-19/Australia/VIC5978/2020\|EPI_ISL_518753\|2020 | .................... | .................... | .................... | .................... | .................... | .................... |

| | | | | | | | |
|---|---|---|---|---|---|---|---|
| G | 4929 hCoV-19/Australia/VIC5979/2020\|EPI_ISL_518754\|2020 | ···················· | ···················· | ····················· | ···················· | ···················· | ···················· |
| G | 4930 hCoV-19/Australia/VIC5980/2020\|EPI_ISL_518755\|2020 | ···················· | ···················· | ····················· | ···················· | ···················· | ···················· |
| G | 4931 hCoV-19/Australia/VIC5981/2020\|EPI_ISL_518756\|2020 | ···················· | ···················· | ····················· | ···················· | ···················· | ···················· |
| G | 4932 hCoV-19/Australia/VIC5982/2020\|EPI_ISL_518757\|2020 | ···················· | ···················· | ····················· | ···················· | ···················· | ···················· |
| G | 4933 hCoV-19/Australia/VIC5983/2020\|EPI_ISL_518758\|2020 | ···················· | ···················· | ····················· | ···················· | ···················· | ···················· |
| G | 4934 hCoV-19/Australia/VIC5984/2020\|EPI_ISL_518759\|2020 | ···················· | ···················· | ····················· | ···················· | ···················· | ···················· |
| G | 4935 hCoV-19/Australia/VIC5985/2020\|EPI_ISL_518760\|2020 | ···················· | ···················· | ····················· | ···················· | ···················· | ···················· |
| G | 4936 hCoV-19/Australia/VIC5986/2020\|EPI_ISL_518761\|2020 | ···················· | ···················· | ····················· | ···················· | ···················· | ···················· |
| G | 4937 hCoV-19/Australia/VIC5987/2020\|EPI_ISL_518762\|2020 | ···················· | ···················· | ····················· | ···················· | ···················· | ···················· |
| G | 4938 hCoV-19/Australia/VIC5988/2020\|EPI_ISL_518763\|2020 | ···················· | ···················· | ····················· | ···················· | ···················· | ···················· |
| G | 4939 hCoV-19/Australia/VIC5990/2020\|EPI_ISL_518765\|2020 | ···················· | ···················· | ····················· | ···················· | ···················· | ···················· |
| G | 4940 hCoV-19/Australia/VIC5991/2020\|EPI_ISL_518766\|2020 | ···················· | ···················· | ····················· | ···················· | ···················· | ···················· |
| G | 4941 hCoV-19/Australia/VIC5992/2020\|EPI_ISL_518767\|2020 | ···················· | ···················· | ····················· | ···················· | ···················· | ···················· |
| G | 4942 hCoV-19/Australia/VIC5993/2020\|EPI_ISL_518768\|2020 | ···················· | ···················· | ····················· | ···················· | ···················· | ···················· |
| G | 4943 hCoV-19/Australia/VIC5994/2020\|EPI_ISL_518769\|2020 | ···················· | ···················· | ····················· | ···················· | ···················· | ···················· |
| G | 4944 hCoV-19/Netherlands/GE-EMC-218/2020\|EPI_ISL_523200 | ···················· | ···················· | ····················· | ···················· | ···················· | ···················· |
| G | 4945 hCoV-19/Netherlands/GE-EMC-220/2020\|EPI_ISL_523202 | ···················· | ···················· | ····················· | ···················· | ···················· | ···················· |
| G | 4946 hCoV-19/Netherlands/GE-EMC-221/2020\|EPI_ISL_523203 | ···················· | ···················· | ····················· | ···················· | ···················· | ···················· |
| G | 4947 hCoV-19/Netherlands/GE-EMC-222/2020\|EPI_ISL_523204 | ···················· | ···················· | ····················· | ···················· | ···················· | ···················· |
| G | 4948 hCoV-19/Netherlands/GE-EMC-224/2020\|EPI_ISL_523205 | ···················· | ···················· | ····················· | ···················· | ···················· | ···················· |
| G | 4949 hCoV-19/Netherlands/GE-EMC-227/2020\|EPI_ISL_523207 | ···················· | ···················· | ····················· | ···················· | ···················· | ···················· |
| G | 4950 hCoV-19/Netherlands/GE-EMC-228/2020\|EPI_ISL_523208 | ···················· | ···················· | ····················· | ···················· | ···················· | ···················· |
| G | 4951 hCoV-19/Netherlands/GE-EMC-229/2020\|EPI_ISL_523209 | ···················· | ···················· | ····················· | ···················· | ···················· | ···················· |
| G | 4952 hCoV-19/Netherlands/GE-EMC-230/2020\|EPI_ISL_523210 | ···················· | ···················· | ····················· | ···················· | ···················· | ···················· |
| G | 4953 hCoV-19/Netherlands/GE-EMC-231/2020\|EPI_ISL_523211 | ···················· | ···················· | ····················· | ···················· | ···················· | ···················· |
| G | 4954 hCoV-19/Netherlands/GE-EMC-234/2020\|EPI_ISL_523213 | ···················· | ···················· | ····················· | ···················· | ···················· | ···················· |
| G | 4955 hCoV-19/Netherlands/GE-EMC-235/2020\|EPI_ISL_523214 | ···················· | ···················· | ····················· | ···················· | ···················· | ···················· |
| G | 4956 hCoV-19/Netherlands/GE-EMC-236/2020\|EPI_ISL_523215 | ···················· | ···················· | ····················· | ···················· | ···················· | ···················· |
| G | 4957 hCoV-19/Netherlands/GE-EMC-237/2020\|EPI_ISL_523216 | ···················· | ···················· | ····················· | ···················· | ···················· | ···················· |
| G | 4958 hCoV-19/Netherlands/GE-EMC-238/2020\|EPI_ISL_523217 | ···················· | ···················· | ····················· | ···················· | ···················· | ···················· |
| G | 4959 hCoV-19/Netherlands/GE-EMC-239/2020\|EPI_ISL_523218 | ···················· | ·A·················· | ····················· | ···················· | ···················· | ··A················· |
| G | 4960 hCoV-19/Netherlands/GE-EMC-241/2020\|EPI_ISL_523220 | ···················· | ···················· | ····················· | ···················· | ···················· | ···················· |
| G | 4961 hCoV-19/Netherlands/GE-EMC-242/2020\|EPI_ISL_523221 | ···················· | ···················· | ····················· | ···················· | ···················· | ···················· |
| G | 4962 hCoV-19/Netherlands/GE-EMC-243/2020\|EPI_ISL_523222 | ···················· | ···················· | ····················· | ···················· | ···················· | ···················· |
| G | 4963 hCoV-19/Netherlands/GE-EMC-244/2020\|EPI_ISL_523223 | ···················· | ···················· | ····················· | ···················· | ···················· | ···················· |
| G | 4964 hCoV-19/Netherlands/GE-EMC-246/2020\|EPI_ISL_523225 | ···················· | ···················· | ····················· | ···················· | ···················· | ···················· |
| G | 4965 hCoV-19/Netherlands/GE-EMC-247/2020\|EPI_ISL_523226 | ···················· | ···················· | ····················· | ···················· | ···················· | ···················· |
| G | 4966 hCoV-19/Netherlands/GE-EMC-248/2020\|EPI_ISL_523227 | ···················· | ···················· | ····················· | ···················· | ···················· | ···················· |
| G | 4967 hCoV-19/Netherlands/GE-EMC-249/2020\|EPI_ISL_523228 | ···················· | ···················· | ····················· | ···················· | ···················· | ···················· |
| G | 4968 hCoV-19/Netherlands/GE-EMC-250/2020\|EPI_ISL_523229 | ···················· | ···················· | ····················· | ···················· | ···················· | ···················· |
| G | 4969 hCoV-19/Netherlands/GE-EMC-251/2020\|EPI_ISL_523230 | ···················· | ···················· | ····················· | ···················· | ···················· | ···················· |
| G | 4970 hCoV-19/Netherlands/GE-EMC-252/2020\|EPI_ISL_523231 | ···················· | ···················· | ····················· | ···················· | ···················· | ···················· |
| G | 4971 hCoV-19/Netherlands/GE-EMC-254/2020\|EPI_ISL_523233 | ···················· | ···················· | ····················· | ···················· | ···················· | ···················· |
| G | 4972 hCoV-19/Netherlands/GE-EMC-256/2020\|EPI_ISL_523235 | ···················· | ···················· | ····················· | ···················· | ···················· | ···················· |
| G | 4973 hCoV-19/Netherlands/GE-EMC-257/2020\|EPI_ISL_523236 | ···················· | ···················· | ····················· | ···················· | ···················· | ···················· |
| G | 4974 hCoV-19/Netherlands/GE-EMC-259/2020\|EPI_ISL_523238 | ···················· | ···················· | ····················· | ···················· | ···················· | ···················· |
| G | 4975 hCoV-19/Netherlands/GE-EMC-260/2020\|EPI_ISL_523239 | ···················· | ···················· | ····················· | ···················· | ···················· | ···················· |
| G | 4976 hCoV-19/Netherlands/GE-EMC-261/2020\|EPI_ISL_523240 | ···················· | ···················· | ····················· | ···················· | ···················· | ···················· |
| G | 4977 hCoV-19/Netherlands/GE-EMC-262/2020\|EPI_ISL_523241 | ···················· | ···················· | ····················· | ···················· | ···················· | ···················· |
| G | 4978 hCoV-19/Netherlands/GE-EMC-266/2020\|EPI_ISL_523245 | ···················· | ···················· | ····················· | ···················· | ···················· | ···················· |
| G | 4979 hCoV-19/Netherlands/GE-EMC-267/2020\|EPI_ISL_523246 | ···················· | ···················· | ····················· | ···················· | ···················· | ···················· |
| G | 4980 hCoV-19/Netherlands/GE-EMC-268/2020\|EPI_ISL_523247 | ···················· | ···················· | ····················· | ···················· | ···················· | ···················· |
| G | 4981 hCoV-19/Netherlands/GE-EMC-269/2020\|EPI_ISL_523248 | ···················· | ···················· | ····················· | ···················· | ···················· | ···················· |
| G | 4982 hCoV-19/Netherlands/GE-EMC-270/2020\|EPI_ISL_523249 | ···················· | ···················· | ····················· | ···················· | ···················· | ···················· |
| G | 4983 hCoV-19/Netherlands/GE-EMC-271/2020\|EPI_ISL_523250 | ···················· | ···················· | ····················· | ···················· | ···················· | ···················· |
| G | 4984 hCoV-19/Netherlands/GE-EMC-272/2020\|EPI_ISL_523251 | ···················· | ···················· | ····················· | ···················· | ···················· | ···················· |
| G | 4985 hCoV-19/Netherlands/GE-EMC-273/2020\|EPI_ISL_523252 | ···················· | ···················· | ····················· | ···················· | ···················· | ···················· |
| G | 4986 hCoV-19/Netherlands/GE-EMC-275/2020\|EPI_ISL_523254 | ···················· | ···················· | ····················· | ···················· | ···················· | ···················· |
| G | 4987 hCoV-19/Netherlands/GE-EMC-278/2020\|EPI_ISL_523257 | ···················· | ···················· | ····················· | ···················· | ···················· | ···················· |
| G | 4988 hCoV-19/Netherlands/GE-EMC-279/2020\|EPI_ISL_523258 | ···················· | ···················· | ····················· | ···················· | ···················· | ···················· |
| G | 4989 hCoV-19/Netherlands/GE-EMC-280/2020\|EPI_ISL_523259 | ···················· | ···················· | ····················· | ···················· | ···················· | ···················· |
| G | 4990 hCoV-19/Netherlands/GE-EMC-281/2020\|EPI_ISL_523260 | ···················· | ···················· | ····················· | ···················· | ···················· | ···················· |
| G | 4991 hCoV-19/Netherlands/GE-EMC-282/2020\|EPI_ISL_523261 | ···················· | ···················· | ····················· | ···················· | ···················· | ···················· |
| G | 4992 hCoV-19/Netherlands/GE-EMC-283/2020\|EPI_ISL_523262 | ···················· | ···················· | ····················· | ···················· | ···················· | ···················· |
| G | 4993 hCoV-19/Netherlands/GE-EMC-284/2020\|EPI_ISL_523263 | ···················· | ···················· | ····················· | ···················· | ···················· | ···················· |
| G | 4994 hCoV-19/Netherlands/GE-EMC-285/2020\|EPI_ISL_523264 | ···················· | ···················· | ····················· | ···················· | ···················· | ···················· |
| G | 4995 hCoV-19/Netherlands/GE-EMC-286/2020\|EPI_ISL_523265 | ···················· | ···················· | ····················· | ···················· | ···················· | ···················· |
| G | 4996 hCoV-19/Netherlands/GE-EMC-288/2020\|EPI_ISL_523267 | ···················· | ···················· | ····················· | ···················· | ···················· | ···················· |
| G | 4997 hCoV-19/Netherlands/GE-EMC-289/2020\|EPI_ISL_523268 | ···················· | ···················· | ····················· | ···················· | ···················· | ···················· |
| G | 4998 hCoV-19/Netherlands/GE-EMC-290/2020\|EPI_ISL_523269 | ···················· | ···················· | ····················· | ···················· | ···················· | ···················· |
| G | 4999 hCoV-19/Netherlands/GE-EMC-291/2020\|EPI_ISL_523270 | ···················· | ···················· | ····················· | ···················· | ···················· | ···················· |
| G | 5000 hCoV-19/Netherlands/GE-EMC-293/2020\|EPI_ISL_523272 | ···················· | ···················· | ····················· | ···················· | ···················· | ···················· |
| G | 5001 hCoV-19/Netherlands/GE-EMC-294/2020\|EPI_ISL_523273 | ···················· | ···················· | ····················· | ···················· | ···················· | ···················· |
| G | 5002 hCoV-19/Netherlands/GE-EMC-295/2020\|EPI_ISL_523274 | ···················· | ···················· | ····················· | ···················· | ···················· | ···················· |
| G | 5003 hCoV-19/Netherlands/GE-EMC-296/2020\|EPI_ISL_523275 | ···················· | ·A·················· | ····················· | ···················· | ···················· | ··A················· |
| G | 5004 hCoV-19/Netherlands/GE-EMC-297/2020\|EPI_ISL_523276 | ···················· | ·A·················· | ····················· | ···················· | ···················· | ··A················· |
| G | 5005 hCoV-19/Netherlands/GE-EMC-298/2020\|EPI_ISL_523277 | ···················· | ·A·················· | ····················· | ···················· | ···················· | ··A················· |
| G | 5006 hCoV-19/Netherlands/GE-EMC-299/2020\|EPI_ISL_523278 | ···················· | ···················· | ····················· | ···················· | ···················· | ···················· |
| G | 5007 hCoV-19/Netherlands/GE-EMC-300/2020\|EPI_ISL_523279 | ···················· | ···················· | ····················· | ···················· | ···················· | ···················· |
| G | 5008 hCoV-19/Netherlands/GE-EMC-301/2020\|EPI_ISL_523280 | ···················· | ···················· | ····················· | ···················· | ···················· | ···················· |

| | | | | | | | | |
|---|---|---|---|---|---|---|---|---|
| G | 5009 hCoV-19/Netherlands/LI-EMC-76/2020\|EPI_ISL_523281\| | ···················· | ···················· | ···················· | ···················· | ···················· | ···················· | |
| G | 5010 hCoV-19/Netherlands/LI-EMC-77/2020\|EPI_ISL_523282\| | ···················· | ···················· | ···················· | ···················· | ···················· | ···················· | |
| G | 5011 hCoV-19/Netherlands/LI-EMC-78/2020\|EPI_ISL_523283\| | ···················· | ···················· | ···················· | ···················· | ···················· | ···················· | |
| G | 5012 hCoV-19/Netherlands/LI-EMC-79/2020\|EPI_ISL_523284\| | ···················· | ···················· | ···················· | ···················· | ···················· | ···················· | |
| G | 5013 hCoV-19/Netherlands/LI-EMC-80/2020\|EPI_ISL_523285\| | ···················· | ···················· | ···················· | ···················· | ···················· | ···················· | |
| G | 5014 hCoV-19/Netherlands/LI-EMC-81/2020\|EPI_ISL_523286\| | ···················· | ···················· | ···················· | ···················· | ···················· | ···················· | |
| G | 5015 hCoV-19/Netherlands/LI-EMC-83/2020\|EPI_ISL_523288\| | ···················· | ···················· | ···················· | ···················· | ···················· | ···················· | |
| G | 5016 hCoV-19/Netherlands/LI-EMC-84/2020\|EPI_ISL_523289\| | ···················· | ···················· | ···················· | ···················· | ···················· | ···················· | |
| G | 5017 hCoV-19/Netherlands/LI-EMC-85/2020\|EPI_ISL_523290\| | ···················· | ···················· | ···················· | ···················· | ···················· | ···················· | |
| G | 5018 hCoV-19/Netherlands/LI-EMC-86/2020\|EPI_ISL_523291\| | ···················· | ···················· | ···················· | ···················· | ···················· | ···················· | |
| G | 5019 hCoV-19/Netherlands/NB-EMC-169/2020\|EPI_ISL_523292 | ···················· | ·A·················· | ···················· | ···················· | ···················· | ··A················· | |
| G | 5020 hCoV-19/Netherlands/NB-EMC-170/2020\|EPI_ISL_523293 | ···················· | ·A·················· | ···················· | ···················· | ···················· | ··A················· | |
| G | 5021 hCoV-19/Netherlands/NB-EMC-171/2020\|EPI_ISL_523294 | ···················· | ·A·················· | ···················· | ···················· | ···················· | ··A················· | |
| G | 5022 hCoV-19/Netherlands/NB-EMC-172/2020\|EPI_ISL_523295 | ···················· | ···················· | ···················· | ···················· | ···················· | ···················· | |
| G | 5023 hCoV-19/Netherlands/NB-EMC-173/2020\|EPI_ISL_523296 | ···················· | ·A·················· | ···················· | ···················· | ···················· | ··A················· | |
| G | 5024 hCoV-19/Netherlands/NB-EMC-174/2020\|EPI_ISL_523297 | ···················· | ·A·················· | ···················· | ···················· | ···················· | ··A················· | |
| G | 5025 hCoV-19/Netherlands/NB-EMC-175/2020\|EPI_ISL_523298 | ···················· | ···················· | ···················· | ···················· | ···················· | ···················· | |
| G | 5026 hCoV-19/Netherlands/NB-EMC-176/2020\|EPI_ISL_523299 | ···················· | ·A·················· | ···················· | ···················· | ···················· | ··A················· | |
| G | 5027 hCoV-19/Scotland/GCVR-1725DC/2020\|EPI_ISL_524526\|2 | ···················· | ···················· | ···················· | ···················· | ···················· | ···················· | |
| G | 5028 hCoV-19/Scotland/GCVR-170D8B/2020\|EPI_ISL_524527\|2 | ···················· | ···················· | ···················· | ···················· | ···················· | ···················· | |
| G | 5029 hCoV-19/England/CAMB-7E401/2020\|EPI_ISL_524530\|202 | ···················· | ···················· | ···················· | ···················· | ···················· | ···················· | |
| G | 5030 hCoV-19/England/CAMB-7E44D/2020\|EPI_ISL_524531\|202 | ···················· | ···················· | ···················· | ···················· | ···················· | ···················· | |
| G | 5031 hCoV-19/Scotland/GCVR-171CC9/2020\|EPI_ISL_524534\|2 | ···················· | ···················A | ···················· | ···················· | ···················· | ···················· | 1 |
| G | 5032 hCoV-19/Scotland/GCVR-1725BE/2020\|EPI_ISL_524535\|2 | ···················· | ···················· | ···················· | ···················· | ···················· | ···················· | |
| G | 5033 hCoV-19/Scotland/GCVR-171E1E/2020\|EPI_ISL_524536\|2 | ···················· | ···················· | ···················· | ···················· | ···················· | ···················· | |
| G | 5034 hCoV-19/England/CAMB-7E53B/2020\|EPI_ISL_524537\|202 | ···················· | ···················· | ···················· | ···················· | ···················· | ···················· | |
| G | 5035 hCoV-19/Scotland/GCVR-171E0F/2020\|EPI_ISL_524539\|2 | ···················· | ···················· | ···················· | ···················· | ···················· | ···················· | |
| G | 5036 hCoV-19/Scotland/GCVR-171BCC/2020\|EPI_ISL_524545\|2 | ···················· | ···················· | ···················· | ···················· | ···················· | ···················· | |
| G | 5037 hCoV-19/Scotland/GCVR-171CE7/2020\|EPI_ISL_524550\|2 | ···················· | ···················A | ···················· | ···················· | ···················· | ···················· | 1 |
| G | 5038 hCoV-19/England/CAMB-80202/2020\|EPI_ISL_524551\|202 | ···················· | ···················· | ···················· | ···················· | ···················· | ···················· | |
| G | 5039 hCoV-19/Scotland/GCVR-1715DD/2020\|EPI_ISL_524552\|2 | ···················· | ···················· | ···················· | ···················· | ···················· | ···················· | |
| G | 5040 hCoV-19/Scotland/GCVR-173034/2020\|EPI_ISL_524556\|2 | ···················· | ···················· | ···················· | ···················· | ···················· | ···················· | |
| G | 5041 hCoV-19/England/CAMB-7E36E/2020\|EPI_ISL_524560\|202 | ···················· | ···················· | ···················· | ···················· | ···················· | ···················· | |
| G | 5042 hCoV-19/Scotland/GCVR-1730DA/2020\|EPI_ISL_524562\|2 | ···················· | ·A·················· | ···················· | ···················· | ···················· | ···················· | |
| G | 5043 hCoV-19/Scotland/GCVR-170E00/2020\|EPI_ISL_524563\|2 | ···················· | ···················· | ···················· | ···················· | ···················· | ···················· | |
| G | 5044 hCoV-19/England/CAMB-802A8/2020\|EPI_ISL_524564\|202 | ···················· | ···················· | ···················· | ···················· | ···················· | ···················· | |
| G | 5045 hCoV-19/Scotland/GCVR-171564/2020\|EPI_ISL_524566\|2 | ···················· | ·············A······ | ···················· | ···················· | ···················· | ···················· | |
| G | 5046 hCoV-19/England/CAMB-7E47A/2020\|EPI_ISL_524568\|202 | ···················· | ···················· | ···················· | ···················· | ···················· | ···················· | |
| G | 5047 hCoV-19/England/CAMB-7F998/2020\|EPI_ISL_524570\|202 | ···················· | ···················· | ···················· | ···················· | ···················· | ···················· | |
| G | 5048 hCoV-19/Scotland/GCVR-173043/2020\|EPI_ISL_524572\|2 | ···················· | ···················· | ···················· | ···················· | ···················· | ···················· | |
| G | 5049 hCoV-19/England/CAMB-1AE777/2020\|EPI_ISL_524573\|20 | ···················· | ···················· | ···················· | ···················· | ···················· | ···················· | |
| G | 5050 hCoV-19/England/CAMB-1AE2C1/2020\|EPI_ISL_524575\|20 | ···················· | ···················· | ···················· | ···················· | ···················· | ···················· | |
| G | 5051 hCoV-19/England/CAMB-1AE6F2/2020\|EPI_ISL_524576\|20 | ···················· | ···················· | ···················· | ···················· | ···················· | ···················· | |
| G | 5052 hCoV-19/England/CAMB-1AE7A4/2020\|EPI_ISL_524577\|20 | ···················· | ···················· | ···················· | ···················· | ···················· | ···················· | |
| G | 5053 hCoV-19/England/CAMB-1AE328/2020\|EPI_ISL_524578\|20 | ···················· | ···················· | ···················· | ···················· | ···················· | ···················· | |
| G | 5054 hCoV-19/England/CAMB-1AE70E/2020\|EPI_ISL_524583\|20 | ···················· | ···················· | ···················· | ···················· | ···················· | ···················· | |
| G | 5055 hCoV-19/England/CAMB-1AEA23/2020\|EPI_ISL_524584\|20 | ···················· | ···················· | ···················· | ···················· | ···················· | ···················· | |
| G | 5056 hCoV-19/England/CAMB-1AE08B/2020\|EPI_ISL_524586\|20 | ···················· | ···················· | ···················· | ···················· | ···················· | ···················· | |
| G | 5057 hCoV-19/England/CAMB-1AE908/2020\|EPI_ISL_524587\|20 | ···················· | ···················· | ···················· | ···················· | ···················· | ···················· | |
| G | 5058 hCoV-19/England/CAMB-1AE5E6/2020\|EPI_ISL_524589\|20 | ···················· | ···················· | ···················· | ···················· | ···················· | ···················· | |
| G | 5059 hCoV-19/England/CAMB-1AE759/2020\|EPI_ISL_524591\|20 | ···················· | ···················· | ···················· | ···················· | ···················· | ···················· | |
| G | 5060 hCoV-19/England/CAMB-1AEAD8/2020\|EPI_ISL_524593\|20 | ···················· | ···················· | ···················· | ···················· | ···················· | ···················· | |
| G | 5061 hCoV-19/England/CAMB-1AE1E2/2020\|EPI_ISL_524594\|20 | ···················· | ···················· | ···················· | ···················· | ···················· | ···················· | |
| G | 5062 hCoV-19/England/CAMB-1AEB3F/2020\|EPI_ISL_524596\|20 | ···················· | ···················· | ···················· | ···················· | ···················· | ···················· | |
| G | 5063 hCoV-19/England/CAMB-1AEAE7/2020\|EPI_ISL_524597\|20 | ···················· | ···················· | ···················· | ···················· | ···················· | ···················· | |
| G | 5064 hCoV-19/England/CAMB-1AE7D1/2020\|EPI_ISL_524598\|20 | ···················· | ···················· | ···················· | ···················· | ···················· | ···················· | |
| G | 5065 hCoV-19/England/CAMB-1AE795/2020\|EPI_ISL_524599\|20 | ···················· | ···················· | ···················· | ···················· | ···················· | ···················· | |
| G | 5066 hCoV-19/USA/OR-OHSU-0631/2020\|EPI_ISL_525857\|2020- | ···················· | ···················· | ···················· | ···················· | ···················· | ···················· | |
| G | 5067 hCoV-19/USA/OR-OHSU-0633/2020\|EPI_ISL_525858\|2020- | ···················· | ···················· | ···················· | ···················· | ···················· | ···················· | |
| G | 5068 hCoV-19/USA/OR-OHSU-0635/2020\|EPI_ISL_525860\|2020- | ···················· | ···················· | ···················· | ···················· | ···················· | ···················· | |
| G | 5069 hCoV-19/USA/OR-OHSU-0637/2020\|EPI_ISL_525862\|2020- | ···················· | ···················· | ···················· | ···················· | ···················· | ···················· | |
| G | 5070 hCoV-19/USA/OR-OHSU-0638/2020\|EPI_ISL_525863\|2020- | ···················· | ···················· | ···················· | ···················· | ···················· | ···················· | |
| G | 5071 hCoV-19/USA/OR-OHSU-0642/2020\|EPI_ISL_525864\|2020- | ···················· | ···················· | ···················· | ···················· | ···················· | ···················· | |
| G | 5072 hCoV-19/USA/OR-OHSU-0646/2020\|EPI_ISL_525867\|2020- | ···················· | ···················· | ···················· | ···················· | ···················· | ···················· | |
| G | 5073 hCoV-19/USA/OR-OHSU-0647/2020\|EPI_ISL_525868\|2020- | ···················· | ···················· | ···················· | ···················· | ···················· | ···················· | |
| G | 5074 hCoV-19/USA/OR-OHSU-0648/2020\|EPI_ISL_525869\|2020- | ···················· | ···················· | ···················· | ···················· | ···················· | ···················· | |
| G | 5075 hCoV-19/USA/OR-OHSU-0649/2020\|EPI_ISL_525870\|2020- | ···················· | ···················· | ···················· | ···················· | ···················· | ···················· | |
| G | 5076 hCoV-19/USA/OR-OHSU-0660/2020\|EPI_ISL_525873\|2020- | ···················· | ···················· | ···················· | ···················· | ···················· | ···················· | |
| G | 5077 hCoV-19/USA/OR-OHSU-0663/2020\|EPI_ISL_525874\|2020- | ···················· | ···················· | ···················· | ···················· | ···················· | ···················· | |
| G | 5078 hCoV-19/USA/OR-OHSU-0665/2020\|EPI_ISL_525876\|2020- | ···················· | ···················· | ···················· | ···················· | ···················· | ···················· | |
| G | 5079 hCoV-19/USA/OR-OHSU-0666/2020\|EPI_ISL_525877\|2020- | ···················· | ·A·················· | ···················· | ···················· | ···················· | ···················· | |
| G | 5080 hCoV-19/USA/OR-OHSU-0667/2020\|EPI_ISL_525878\|2020- | ···················· | ···················· | ···················· | ···················· | ···················· | ···················· | |
| G | 5081 hCoV-19/USA/OR-OHSU-0671/2020\|EPI_ISL_525880\|2020- | ···················· | ···················· | ···················· | ···················· | ···················· | ···················· | |
| G | 5082 hCoV-19/USA/OR-OHSU-0673/2020\|EPI_ISL_525881\|2020- | ···················· | ···················· | ···················· | ···················· | ···················· | ···················· | |
| G | 5083 hCoV-19/USA/OR-OHSU-0675/2020\|EPI_ISL_525883\|2020- | ···················· | ···················· | ···················· | ···················· | ···················· | ···················· | |
| G | 5084 hCoV-19/USA/OR-OHSU-0683/2020\|EPI_ISL_525885\|2020- | ···················· | ·A·················· | ···················· | ···················· | ···················· | ···················· | |
| G | 5085 hCoV-19/USA/OR-OHSU-0713/2020\|EPI_ISL_525891\|2020- | ···················· | ·A·················· | ···················· | ···················· | ···················· | ···················· | |
| G | 5086 hCoV-19/USA/OR-OHSU-0721/2020\|EPI_ISL_525892\|2020- | ···················· | ···················· | ···················· | ···················· | ···················· | ···················· | |
| G | 5087 hCoV-19/USA/OR-OHSU-0725/2020\|EPI_ISL_525893\|2020- | ···················· | ···················· | ···················· | ···················· | ···················· | ···················· | |
| G | 5088 hCoV-19/USA/OR-OHSU-0767/2020\|EPI_ISL_525894\|2020- | ···················· | ···················· | ···················· | ···················· | ···················· | ···················· | |

| | | | | | | | | |
|---|---|---|---|---|---|---|---|---|
| G | 5089 | hCoV-19/USA/OR-OHSU-0771/2020\|EPI_ISL_525895\|2020- | ···················· | ·A·················· | ····················· | ···················· | ···················· | ···················· |
| G | 5090 | hCoV-19/USA/OR-OHSU-0773/2020\|EPI_ISL_525896\|2020- | ···················· | ···················· | ····················· | ···················· | ···················· | ···················· |
| G | 5091 | hCoV-19/USA/OR-OHSU-0775/2020\|EPI_ISL_525897\|2020- | ···················· | ···················· | ····················· | ···················· | ···················· | ···················· |
| G | 5092 | hCoV-19/USA/OR-OHSU-0777/2020\|EPI_ISL_525898\|2020- | ···················· | ···················· | ····················· | ···················· | ···················· | ···················· |
| G | 5093 | hCoV-19/USA/OR-OHSU-0783/2020\|EPI_ISL_525899\|2020- | ···················· | ···················· | ····················· | ···················· | ···················· | ···················· |
| G | 5094 | hCoV-19/Switzerland/BL-UHB-42189772/2020\|EPI_ISL_5 | ···················· | ···················· | ····················· | ···················· | ···················· | ···················· |
| G | 5095 | hCoV-19/Switzerland/BL-UHB-42189773/2020\|EPI_ISL_5 | ···················· | ···················· | ····················· | ···················· | ···················· | ···················· |
| G | 5096 | hCoV-19/Switzerland/BS-UHB-42189793/2020\|EPI_ISL_5 | ···················· | ···················· | ····················· | ···················· | ···················· | ···················· |
| G | 5097 | hCoV-19/Switzerland/BS-UHB-42189861/2020\|EPI_ISL_5 | ···················· | ···················· | ····················· | ···················· | ···················· | ···················· |
| G | 5098 | hCoV-19/Switzerland/BS-UHB-42189862/2020\|EPI_ISL_5 | ···················· | ···················· | ····················· | ···················· | ···················· | ···················· |
| G | 5099 | hCoV-19/Switzerland/BS-UHB-42189866/2020\|EPI_ISL_5 | ···················· | ···················· | ····················· | ···················· | ···················· | ···················· |
| G | 5100 | hCoV-19/Switzerland/VD-UHB-42192980/2020\|EPI_ISL_5 | ···················· | ···················· | ····················· | ···················· | ···················· | ···················· |
| G | 5101 | hCoV-19/Switzerland/BS-UHB-42193014/2020\|EPI_ISL_5 | ···················· | ···················· | ····················· | ···················· | ···················· | ···················· |
| G | 5102 | hCoV-19/Switzerland/BS-UHB-42193056/2020\|EPI_ISL_5 | ···················· | ·A·················· | ····················· | ···················· | ···················· | ···················· |
| G | 5103 | hCoV-19/Switzerland/BS-UHB-42193080/2020\|EPI_ISL_5 | ···················· | ···················· | ····················· | ···················· | ···················· | ···················· |
| G | 5104 | hCoV-19/Switzerland/BL-UHB-42193250/2020\|EPI_ISL_5 | ···················· | ···················· | ····················· | ···················· | ···················· | ···················· |
| G | 5105 | hCoV-19/Switzerland/BL-UHB-42193254/2020\|EPI_ISL_5 | ···················· | ···················· | ····················· | ···················· | ···················· | ···················· |
| G | 5106 | hCoV-19/Switzerland/BS-UHB-42193283/2020\|EPI_ISL_5 | ···················· | ···················· | ····················· | ···················· | ···················· | ···················· |
| G | 5107 | hCoV-19/Switzerland/BS-UHB-42193328/2020\|EPI_ISL_5 | ···················· | ···················· | ····················· | ···················· | ···················· | ···················· |
| G | 5108 | hCoV-19/Switzerland/BS-UHB-42193780/2020\|EPI_ISL_5 | ···················· | ···················· | ····················· | ···················· | ···················· | ···················· |
| G | 5109 | hCoV-19/Switzerland/BL-UHB-42194077/2020\|EPI_ISL_5 | ···················· | ···················· | ····················· | ···················· | ···················· | ···················· |
| G | 5110 | hCoV-19/Switzerland/BL-UHB-42194080/2020\|EPI_ISL_5 | ···················· | ···················· | ····················· | ···················· | ···················· | ···················· |
| G | 5111 | hCoV-19/Switzerland/BL-UHB-42194287/2020\|EPI_ISL_5 | ···················· | ···················· | ····················· | ···················· | ···················· | ···················· |
| G | 5112 | hCoV-19/Switzerland/BL-UHB-42194288/2020\|EPI_ISL_5 | ···················· | ···················· | ····················· | ···················· | ···················· | ···················· |
| G | 5113 | hCoV-19/Switzerland/BL-UHB-42194515/2020\|EPI_ISL_5 | ···················· | ···················· | ····················· | ···················· | ···················· | ···················· |
| G | 5114 | hCoV-19/Switzerland/BS-UHB-42194519/2020\|EPI_ISL_5 | ···················· | ···················· | ····················· | ···················· | ···················· | ···················· |
| G | 5115 | hCoV-19/Switzerland/BS-UHB-42194551/2020\|EPI_ISL_5 | ···················· | ···················· | ····················· | ···················· | ···················· | ···················· |
| G | 5116 | hCoV-19/Switzerland/BS-UHB-42194578/2020\|EPI_ISL_5 | ···················· | ···················· | ····················· | ···················· | ···················· | ···················· |
| G | 5117 | hCoV-19/Switzerland/BS-UHB-42194826/2020\|EPI_ISL_5 | ···················· | ···················· | ····················· | ···················· | ···················· | ···················· |
| G | 5118 | hCoV-19/Switzerland/BS-UHB-42194901/2020\|EPI_ISL_5 | ···················· | ···················· | ····················· | ···················· | ···················· | ···················· |
| G | 5119 | hCoV-19/Switzerland/BS-UHB-42194904/2020\|EPI_ISL_5 | ···················· | ···················· | ····················· | ···················· | ···················· | ···················· |
| G | 5120 | hCoV-19/Switzerland/BL-UHB-42194939/2020\|EPI_ISL_5 | ···················· | ···················· | ····················· | ···················· | ···················· | ···················· |
| G | 5121 | hCoV-19/England/LIVE-A924F/2020\|EPI_ISL_517281\|202 | ···················· | ···················· | ····················· | ···················· | ···················· | ···················· |
| G | 5124 | hCoV-19/England/LIVE-A929A/2020\|EPI_ISL_517286\|202 | ···················· | ···················· | ····················· | ···················· | ···················· | ···················· |
| G | 5125 | hCoV-19/England/LIVE-A92A9/2020\|EPI_ISL_517287\|202 | ···················· | ···················· | ····················· | ···················· | ···················· | ···················· |
| G | 5126 | hCoV-19/England/LIVE-A92B8/2020\|EPI_ISL_517288\|202 | ···················· | ···················· | ····················· | ···················· | ···················· | ···················· |
| G | 5127 | hCoV-19/England/LIVE-A92C7/2020\|EPI_ISL_517289\|202 | ···················· | ···················· | ····················· | ···················· | ···················· | ···················· |
| G | 5129 | hCoV-19/England/LIVE-A92F4/2020\|EPI_ISL_517292\|202 | ···················· | ·A·················· | ····················· | ···················· | ···················· | ···················· |
| G | 5130 | hCoV-19/England/LIVE-A9300/2020\|EPI_ISL_517293\|202 | ···················· | ···················· | ····················· | ···················· | ···················· | ···················· |
| G | 5131 | hCoV-19/England/LIVE-A993B/2020\|EPI_ISL_517294\|202 | ···················· | ···················· | ····················· | ···················· | ···················· | ···················· |
| G | 5132 | hCoV-19/England/LIVE-A9959/2020\|EPI_ISL_517296\|202 | ···················· | ···················· | ····················· | ···················· | ···················· | ···················· |
| G | 5133 | hCoV-19/England/LIVE-A9968/2020\|EPI_ISL_517297\|202 | ···················· | ···················· | ····················· | ···················· | ···················· | ···················· |
| G | 5134 | hCoV-19/England/LIVE-AA3A5/2020\|EPI_ISL_517444\|202 | ···················· | ···················· | ····················· | ···················· | ···················· | ···················· |
| G | 5135 | hCoV-19/England/LIVE-AA3E1/2020\|EPI_ISL_517448\|202 | ···················· | ···················· | ····················· | ···················· | ···················· | ···················· |
| G | 5136 | hCoV-19/England/LIVE-AA3F0/2020\|EPI_ISL_517449\|202 | ···················· | ···················· | ····················· | ···················· | ···················· | ···················· |
| G | 5137 | hCoV-19/England/LIVE-AA42A/2020\|EPI_ISL_517452\|202 | ···················· | ···················· | ····················· | ···················· | ···················· | ···················· |
| G | 5141 | hCoV-19/England/LIVE-AA5AF/2020\|EPI_ISL_517469\|202 | ···················· | ···················· | ····················· | ···················· | ···················· | ···················· |
| G | 5142 | hCoV-19/England/LIVE-AA5BE/2020\|EPI_ISL_517470\|202 | ···················· | ···················· | ····················· | ···················· | ···················· | ···················· |
| G | 5143 | hCoV-19/England/LIVE-AA5CD/2020\|EPI_ISL_517471\|202 | ···················· | ···················· | ····················· | ···················· | ···················· | ···················· |
| G | 5144 | hCoV-19/England/LIVE-AA5DC/2020\|EPI_ISL_517472\|202 | ···················· | ···················· | ····················· | ···················· | ···················· | ···················· |
| G | 5145 | hCoV-19/England/LIVE-AA5EB/2020\|EPI_ISL_517473\|202 | ···················· | ···················· | ····················· | ···················· | ···················· | ···················· |
| G | 5146 | hCoV-19/England/LIVE-AA606/2020\|EPI_ISL_517475\|202 | ···················· | ···················· | ····················· | ···················· | ···················· | ···················· |
| G | 5147 | hCoV-19/England/LIVE-AA615/2020\|EPI_ISL_517476\|202 | ···················· | ···················· | ····················· | ···················· | ···················· | ···················· |
| G | 5148 | hCoV-19/England/LIVE-AA624/2020\|EPI_ISL_517477\|202 | ···················· | ···················· | ····················· | ···················· | ···················· | ···················· |
| G | 5150 | hCoV-19/England/LIVE-AA651/2020\|EPI_ISL_517480\|202 | ···················· | ···················· | ····················· | ···················· | ···················· | ···················· |
| G | 5152 | hCoV-19/England/LIVE-AA67F/2020\|EPI_ISL_517482\|202 | ···················· | ···················· | ····················· | ···················· | ···················· | ···················· |
| G | 5153 | hCoV-19/England/LIVE-AA68E/2020\|EPI_ISL_517483\|202 | ···················· | ···················· | ····················· | ···················· | ···················· | ···················· |
| G | 5154 | hCoV-19/England/LIVE-AA69D/2020\|EPI_ISL_517484\|202 | ···················· | ···················· | ····················· | ···················· | ···················· | ···················· |
| G | 5155 | hCoV-19/England/LIVE-AA6AC/2020\|EPI_ISL_517485\|202 | ···················· | ···················· | ····················· | ···················· | ···················· | ···················· |
| G | 5156 | hCoV-19/England/LIVE-AA6BB/2020\|EPI_ISL_517486\|202 | ···················· | ···················· | ····················· | ···················· | ···················· | ···················· |
| G | 5157 | hCoV-19/England/LIVE-AA6CA/2020\|EPI_ISL_517487\|202 | ···················· | ···················· | ····················· | ···················· | ···················· | ···················· |
| G | 5158 | hCoV-19/England/LIVE-AA6D9/2020\|EPI_ISL_517488\|202 | ···················· | ···················· | ····················· | ···················· | ···················· | ···················· |
| G | 5159 | hCoV-19/England/LIVE-AA6E8/2020\|EPI_ISL_517489\|202 | ···················· | ···················· | ····················· | ···················· | ···················· | ···················· |
| G | 5160 | hCoV-19/England/LIVE-AA712/2020\|EPI_ISL_517492\|202 | ···················· | ···················· | ····················· | ···················· | ···················· | ···················· |
| G | 5161 | hCoV-19/England/LIVE-AA721/2020\|EPI_ISL_517493\|202 | ···················· | ···················· | ····················· | ···················· | ···················· | ···················· |
| G | 5162 | hCoV-19/England/LIVE-AA730/2020\|EPI_ISL_517494\|202 | ···················· | ···················· | ····················· | ···················· | ···················· | ···················· |
| G | 5164 | hCoV-19/Australia/VIC5995/2020\|EPI_ISL_518770\|2020 | ···················· | ···················· | ····················· | ···················· | ···················· | ···················· |
| G | 5165 | hCoV-19/Australia/VIC5996/2020\|EPI_ISL_518771\|2020 | ···················· | ···················· | ····················· | ···················· | ···················· | ···················· |
| G | 5166 | hCoV-19/Australia/VIC5998/2020\|EPI_ISL_518773\|2020 | ···················· | ···················· | ····················· | ···················· | ···················· | ···················· |
| G | 5167 | hCoV-19/Australia/VIC5999/2020\|EPI_ISL_518774\|2020 | ···················· | ···················· | ····················· | ···················· | ···················· | ···················· |
| G | 5168 | hCoV-19/Australia/VIC6000/2020\|EPI_ISL_518775\|2020 | ···················· | ···················· | ····················· | ···················· | ···················· | ···················· |
| G | 5169 | hCoV-19/Australia/VIC6001/2020\|EPI_ISL_518776\|2020 | ···················· | ···················· | ····················· | ···················· | ···················· | ···················· |
| G | 5170 | hCoV-19/Australia/VIC6002/2020\|EPI_ISL_518777\|2020 | ···················· | ··········A········· | ····················· | ···················· | ···················· | ···················· |
| G | 5171 | hCoV-19/Australia/VIC6003/2020\|EPI_ISL_518778\|2020 | ···················· | ···················· | ····················· | ···················· | ···················· | ···················· |
| G | 5172 | hCoV-19/Australia/VIC6004/2020\|EPI_ISL_518779\|2020 | ···················· | ···················· | ····················· | ···················· | ···················· | ···················· |
| G | 5173 | hCoV-19/Australia/VIC6005/2020\|EPI_ISL_518780\|2020 | ···················· | ···················· | ····················· | ···················· | ···················· | ···················· |
| G | 5174 | hCoV-19/Australia/VIC6006/2020\|EPI_ISL_518781\|2020 | ···················· | ···················· | ····················· | ···················· | ···················· | ···················· |
| G | 5175 | hCoV-19/Australia/VIC6007/2020\|EPI_ISL_518782\|2020 | ···················· | ···················· | ····················· | ···················· | ···················· | ···················· |
| G | 5176 | hCoV-19/Australia/VIC6008/2020\|EPI_ISL_518783\|2020 | ···················· | ···················· | ····················· | ···················· | ···················· | ···················· |
| G | 5177 | hCoV-19/Australia/VIC6009/2020\|EPI_ISL_518784\|2020 | ···················· | ···················· | ····················· | ···················· | ···················· | ···················· |

| | | | | | | | | |
|---|---|---|---|---|---|---|---|---|
| G | 5178 | hCoV-19/Australia/VIC6010/2020\|EPI_ISL_518785\|2020 | .................... | .................... | ..................... | .................... | .................... | .................... |
| G | 5179 | hCoV-19/Australia/VIC6011/2020\|EPI_ISL_518786\|2020 | .................... | .................... | ..................... | .................... | .................... | .................... |
| G | 5180 | hCoV-19/Australia/VIC6012/2020\|EPI_ISL_518787\|2020 | .................... | .................... | ..................... | .................... | .................... | .................... |
| G | 5181 | hCoV-19/Australia/VIC6013/2020\|EPI_ISL_518788\|2020 | .................... | .................... | ..................... | .................... | .................... | .................... |
| G | 5182 | hCoV-19/Australia/VIC6014/2020\|EPI_ISL_518789\|2020 | .................... | .................... | ..................... | .................... | .................... | .................... |
| G | 5183 | hCoV-19/Australia/VIC6015/2020\|EPI_ISL_518790\|2020 | .................... | .................... | ..................... | .................... | .................... | .................... |
| G | 5184 | hCoV-19/Australia/VIC6016/2020\|EPI_ISL_518791\|2020 | .................... | .................... | ..................... | .................... | .................... | .................... |
| G | 5185 | hCoV-19/Australia/VIC6017/2020\|EPI_ISL_518792\|2020 | .................... | .................... | ..................... | .................... | .................... | .................... |
| G | 5186 | hCoV-19/Australia/VIC6020/2020\|EPI_ISL_518795\|2020 | .................... | .................... | ..................... | .................... | .................... | .................... |
| G | 5187 | hCoV-19/Australia/VIC6021/2020\|EPI_ISL_518796\|2020 | .................... | .................... | ..................... | .................... | .................... | .................... |
| G | 5188 | hCoV-19/Suriname/SR-50/2020\|EPI_ISL_518799\|2020-07 | .................... | .................... | ..................... | .................... | .................... | .................... |
| G | 5189 | hCoV-19/mink/Netherlands/NB-EMC-6-15/2020\|EPI_ISL_ | .................... | .................... | ..................... | .................... | .................... | .................... |
| G | 5190 | hCoV-19/mink/Netherlands/NB-EMC-6-16/2020\|EPI_ISL_ | .................... | .................... | ..................... | .................... | .................... | .................... |
| G | 5191 | hCoV-19/mink/Netherlands/NB-EMC-6-18/2020\|EPI_ISL_ | .................... | .................... | ..................... | .................... | .................... | .................... |
| G | 5192 | hCoV-19/mink/Netherlands/NB-EMC-6-2/2020\|EPI_ISL_5 | .................... | .................... | ..................... | .................... | .................... | .................... |
| G | 5193 | hCoV-19/mink/Netherlands/NB-EMC-6-3/2020\|EPI_ISL_5 | .................... | .................... | ..................... | .................... | .................... | .................... |
| G | 5194 | hCoV-19/mink/Netherlands/NB-EMC-6-7/2020\|EPI_ISL_5 | .................... | .................... | ..................... | .................... | .................... | .................... |
| G | 5195 | hCoV-19/mink/Netherlands/NB-EMC-7-10/2020\|EPI_ISL_ | .................... | .................... | ..................... | .................... | .................... | .................... |
| G | 5196 | hCoV-19/mink/Netherlands/NB-EMC-7-19/2020\|EPI_ISL_ | .................... | .................... | ..................... | .................... | .................... | .................... |
| G | 5197 | hCoV-19/mink/Netherlands/NB-EMC-7-2/2020\|EPI_ISL_5 | .................... | .................... | ..................... | .................... | .................... | .................... |
| G | 5198 | hCoV-19/mink/Netherlands/NB-EMC-7-3/2020\|EPI_ISL_5 | .................... | .................... | ..................... | .................... | .................... | .................... |
| G | 5199 | hCoV-19/mink/Netherlands/NB-EMC-7-6/2020\|EPI_ISL_5 | .................... | .................... | ..................... | .................... | .................... | .................... |
| G | 5200 | hCoV-19/mink/Netherlands/NB-EMC-8-11/2020\|EPI_ISL_ | .................... | .................... | ..................... | .................... | .................... | .................... |
| G | 5201 | hCoV-19/mink/Netherlands/NB-EMC-8-14/2020\|EPI_ISL_ | .................... | .................... | ..................... | .................... | .................... | .................... |
| G | 5202 | hCoV-19/mink/Netherlands/NB-EMC-8-20/2020\|EPI_ISL_ | .................... | .................... | ..................... | .................... | .................... | .................... |
| G | 5203 | hCoV-19/mink/Netherlands/NB-EMC-8-8/2020\|EPI_ISL_5 | .................... | .................... | ..................... | .................... | .................... | .................... |
| G | 5204 | hCoV-19/mink/Netherlands/NB-EMC-8-9/2020\|EPI_ISL_5 | .................... | .................... | ..................... | .................... | .................... | .................... |
| G | 5205 | hCoV-19/mink/Netherlands/NB-EMC-9-1/2020\|EPI_ISL_5 | .................... | .................... | ..................... | .................... | .................... | .................... |
| G | 5206 | hCoV-19/Netherlands/DR-EMC-10/2020\|EPI_ISL_523121\| | .................... | .................... | ..................... | .................... | .................... | .................... |
| G | 5207 | hCoV-19/Netherlands/DR-EMC-4/2020\|EPI_ISL_523122\|2 | .................... | .................... | ..................... | .................... | .................... | .................... |
| G | 5208 | hCoV-19/Netherlands/DR-EMC-5/2020\|EPI_ISL_523123\|2 | .................... | .................... | ..................... | .................... | .................... | .................... |
| G | 5209 | hCoV-19/Netherlands/DR-EMC-6/2020\|EPI_ISL_523124\|2 | .................... | .................... | ..................... | .................... | .................... | .................... |
| G | 5210 | hCoV-19/Netherlands/DR-EMC-7/2020\|EPI_ISL_523125\|2 | .................... | .................... | ..................... | .................... | .................... | .................... |
| G | 5211 | hCoV-19/Netherlands/DR-EMC-8/2020\|EPI_ISL_523126\|2 | .................... | .................... | ..................... | .................... | .................... | .................... |
| G | 5212 | hCoV-19/Netherlands/DR-EMC-9/2020\|EPI_ISL_523127\|2 | .................... | .................... | ..................... | .................... | .................... | .................... |
| G | 5213 | hCoV-19/Netherlands/FL-EMC-19/2020\|EPI_ISL_523128\| | .................... | .................... | ..................... | .................... | .................... | .................... |
| G | 5214 | hCoV-19/Netherlands/FL-EMC-20/2020\|EPI_ISL_523129\| | .................... | .................... | ..................... | .................... | .................... | .................... |
| G | 5215 | hCoV-19/Netherlands/FL-EMC-21/2020\|EPI_ISL_523130\| | .................... | .................... | ..................... | .................... | .................... | .................... |
| G | 5216 | hCoV-19/Netherlands/FL-EMC-23/2020\|EPI_ISL_523132\| | .................... | .................... | ..................... | .................... | .................... | .................... |
| G | 5217 | hCoV-19/Netherlands/FL-EMC-24/2020\|EPI_ISL_523133\| | .................... | .................... | ..................... | .................... | .................... | .................... |
| G | 5218 | hCoV-19/Netherlands/FL-EMC-25/2020\|EPI_ISL_523134\| | .................... | .................... | ..................... | .................... | .................... | .................... |
| G | 5219 | hCoV-19/Netherlands/FR-EMC-46/2020\|EPI_ISL_523138\| | .................... | .................... | ..................... | .................... | .................... | .................... |
| G | 5220 | hCoV-19/Netherlands/FR-EMC-47/2020\|EPI_ISL_523139\| | .................... | .................... | ..................... | .................... | .................... | .................... |
| G | 5221 | hCoV-19/Netherlands/FR-EMC-48/2020\|EPI_ISL_523140\| | .................... | .................... | ..................... | .................... | .................... | .................... |
| G | 5222 | hCoV-19/Netherlands/FR-EMC-49/2020\|EPI_ISL_523141\| | .................... | .................... | ..................... | .................... | .................... | .................... |
| G | 5223 | hCoV-19/Netherlands/FR-EMC-50/2020\|EPI_ISL_523142\| | .................... | .................... | ..................... | .................... | .................... | .................... |
| G | 5224 | hCoV-19/Netherlands/FR-EMC-51/2020\|EPI_ISL_523143\| | .................... | .................... | ..................... | .................... | .................... | .................... |
| G | 5225 | hCoV-19/Netherlands/FR-EMC-52/2020\|EPI_ISL_523144\| | .................... | .................... | ..................... | .................... | .................... | .................... |
| G | 5226 | hCoV-19/Netherlands/FR-EMC-53/2020\|EPI_ISL_523145\| | .................... | .................... | ..................... | .................... | .................... | .................... |
| G | 5227 | hCoV-19/Netherlands/FR-EMC-54/2020\|EPI_ISL_523146\| | .................... | .................... | ..................... | .................... | .................... | .................... |
| G | 5228 | hCoV-19/Netherlands/GE-EMC-164/2020\|EPI_ISL_523147 | .................... | .................... | ..................... | .................... | .................... | .................... |
| G | 5229 | hCoV-19/Netherlands/GE-EMC-165/2020\|EPI_ISL_523148 | .................... | .................... | ..................... | .................... | .................... | .................... |
| G | 5230 | hCoV-19/Netherlands/GE-EMC-166/2020\|EPI_ISL_523149 | .................... | .................... | ..................... | .................... | .................... | .................... |
| G | 5231 | hCoV-19/Netherlands/GE-EMC-167/2020\|EPI_ISL_523150 | .................... | .................... | ..................... | .................... | .................... | .................... |
| G | 5232 | hCoV-19/Netherlands/GE-EMC-168/2020\|EPI_ISL_523151 | .................... | .................... | ..................... | .................... | .................... | .................... |
| G | 5233 | hCoV-19/Netherlands/GE-EMC-169/2020\|EPI_ISL_523152 | .................... | .................... | ..................... | .................... | .................... | .................... |
| G | 5234 | hCoV-19/Netherlands/GE-EMC-170/2020\|EPI_ISL_523153 | .................... | .................... | ..................... | .................... | .................... | .................... |
| G | 5235 | hCoV-19/Netherlands/GE-EMC-171/2020\|EPI_ISL_523154 | .................... | .................... | ..................... | .................... | .................... | .................... |
| G | 5236 | hCoV-19/Netherlands/GE-EMC-173/2020\|EPI_ISL_523156 | .................... | .................... | ..................... | .................... | .................... | .................... |
| G | 5237 | hCoV-19/Netherlands/GE-EMC-175/2020\|EPI_ISL_523157 | .................... | .................... | ..................... | .................... | .................... | .................... |
| G | 5238 | hCoV-19/Netherlands/GE-EMC-176/2020\|EPI_ISL_523158 | .................... | .................... | ..................... | .................... | .................... | .................... |
| G | 5239 | hCoV-19/Netherlands/GE-EMC-177/2020\|EPI_ISL_523159 | .................... | .................... | ..................... | .................... | .................... | .................... |
| G | 5240 | hCoV-19/USA/UT-08677/2020\|EPI_ISL_524400\|2020-05-2 | .................... | .................... | ..................... | .................... | .................... | .................... |
| G | 5241 | hCoV-19/USA/UT-08678/2020\|EPI_ISL_524401\|2020-05-2 | .................... | .................... | ..................... | .................... | .................... | .................... |
| G | 5242 | hCoV-19/USA/UT-08680/2020\|EPI_ISL_524402\|2020-05-0 | .................... | .................... | ..................... | .................... | .................... | .................... |
| G | 5243 | hCoV-19/USA/UT-08681/2020\|EPI_ISL_524403\|2020-05-0 | .................... | .................... | ..................... | .................... | .................... | .................... |
| G | 5244 | hCoV-19/USA/UT-08683/2020\|EPI_ISL_524404\|2020-05-0 | .................... | .................... | ..................... | .................... | .................... | .................... |
| G | 5245 | hCoV-19/USA/UT-08693/2020\|EPI_ISL_524412\|2020-05-0 | .................... | .................... | ..................... | .................... | .................... | .................... |
| G | 5246 | hCoV-19/USA/UT-08695/2020\|EPI_ISL_524413\|2020-05-2 | .................... | .................... | ..................... | .................... | .................... | .................... |
| G | 5247 | hCoV-19/USA/UT-08696/2020\|EPI_ISL_524414\|2020-05-2 | .................... | .................... | ..................... | .................... | .................... | .................... |
| G | 5248 | hCoV-19/USA/UT-08697/2020\|EPI_ISL_524415\|2020-05-1 | .................... | .................... | ..................... | .................... | .................... | .................... |
| G | 5249 | hCoV-19/USA/UT-08698/2020\|EPI_ISL_524416\|2020-05-0 | .................... | .................... | ..................... | .................... | .................... | .................... |
| G | 5250 | hCoV-19/USA/UT-08699/2020\|EPI_ISL_524417\|2020-05-0 | .................... | .................... | ..................... | .................... | .................... | .................... |
| G | 5251 | hCoV-19/USA/UT-08700/2020\|EPI_ISL_524418\|2020-05-0 | .................... | .................... | ..................... | .................... | .................... | .................... |
| G | 5252 | hCoV-19/USA/UT-08702/2020\|EPI_ISL_524419\|2020-05-0 | .................... | .................... | ..................... | .................... | .................... | .................... |
| G | 5253 | hCoV-19/USA/UT-08703/2020\|EPI_ISL_524420\|2020-05-0 | .................... | .................... | ..................... | .................... | .................... | .................... |
| G | 5254 | hCoV-19/USA/UT-08704/2020\|EPI_ISL_524421\|2020-05-2 | .................... | .................... | ..................... | .................... | .................... | .................... |
| G | 5255 | hCoV-19/USA/UT-08707/2020\|EPI_ISL_524424\|2020-05-2 | .................... | .................... | ..................... | .................... | .................... | .................... |
| G | 5256 | hCoV-19/USA/UT-08708/2020\|EPI_ISL_524425\|2020-05-2 | .................... | .................... | ..................... | .................... | .................... | .................... |
| G | 5257 | hCoV-19/USA/NC-200414/2020\|EPI_ISL_524430\|2020-03- | .................... | .................... | ..................... | .................... | .................... | .................... |

G 5258 hCoV-19/Singapore/743/2020|EPI_ISL_524445|2020-07- ···················· ···················· ····················· ···················· ···················· ····················
G 5259 hCoV-19/Singapore/740/2020|EPI_ISL_524447|2020-07- ···················· ···················· ····················· ···················· ···················· ····················
G 5260 hCoV-19/Singapore/741/2020|EPI_ISL_524448|2020-07- ···················· ···················· ····················· ···················· ···················· ····················
G 5261 hCoV-19/Singapore/786/2020|EPI_ISL_524449|2020-06- ···················· ···················· ····················· ···················· ···················· ····················
G 5262 hCoV-19/Brazil/SP-272/2020|EPI_ISL_524462|2020-03- ···················· ···················· ····················· ···················· ···················· ····················
G 5263 hCoV-19/Brazil/SP-275/2020|EPI_ISL_524463|2020-03- ···················· ···················· ····················· ···················· ···················· ····················
G 5264 hCoV-19/Brazil/SP-276/2020|EPI_ISL_524464|2020-03- ···················· ···················· ····················· ···················· ···················· ····················
G 5265 hCoV-19/Brazil/SP-511/2020|EPI_ISL_524465|2020-04- ···················· ···················· ····················· ···················· ···················· ····················
G 5266 hCoV-19/Brazil/SP-262/2020|EPI_ISL_524466|2020-04- ···················· ···················· ····················· ···················· ···················· ····················
G 5267 hCoV-19/Brazil/SP-263/2020|EPI_ISL_524467|2020-04- ···················· ···················· ····················· ···················· ···················· ····················
G 5268 hCoV-19/Brazil/SP-240/2020|EPI_ISL_524468|2020-05- ···················· ···················· ····················· ···················· ···················· ····················
G 5269 hCoV-19/Brazil/SP-254/2020|EPI_ISL_524469|2020-05- ···················· ···················· ····················· ···················· ···················· ····················
G 5270 hCoV-19/Brazil/SP-259/2020|EPI_ISL_524470|2020-05- ···················· ···················· ····················· ···················· ···················· ····················
G 5271 hCoV-19/Peru/JUN-INS-074/2020|EPI_ISL_524471|2020- ···················· ···················· ····················· ···················· ···················· ····················
G 5272 hCoV-19/Peru/LIM-INS-060/2020|EPI_ISL_524473|2020- ···················· ···················· ····················· ···················· ···················· ····················
G 5273 hCoV-19/Switzerland/AG-UZH-1000477914/2020|EPI_ISL ···················· ···················· ····················· ···················· ···················· ····················
G 5274 hCoV-19/Switzerland/ZH-UZH-1000477916/2020|EPI_ISL ···················· ···················· ····················· ···················· ···················· ····················
G 5275 hCoV-19/Switzerland/AG-UZH-1000478260/2020|EPI_ISL ···················· ···················· ····················· ···················· ···················· ····················
G 5276 hCoV-19/Switzerland/ZH-UZH-1000478309/2020|EPI_ISL ···················· ···················· ····················· ···················· ···················· ····················
G 5277 hCoV-19/Switzerland/ZH-UZH-1000478320/2020|EPI_ISL ···················· ···················· ····················· ···················· ···················· ····················
G 5278 hCoV-19/Switzerland/ZH-UZH-1000478554/2020|EPI_ISL ···················· ···················· ····················· ···················· ···················· ····················
G 5279 hCoV-19/Switzerland/AG-UZH-1000478635/2020|EPI_ISL ···················· ···················· ····················· ···················· ···················· ····················
G 5280 hCoV-19/Switzerland/AG-UZH-1000478772/2020|EPI_ISL ···················· ···················· ····················· ···················· ···················· ····················
G 5281 hCoV-19/Switzerland/ZH-UZH-1000478518/2020|EPI_ISL ···················· ···················· ····················· ···················· ···················· ····················
G 5282 hCoV-19/Switzerland/ZH-UZH-1000478869/2020|EPI_ISL ···················· ···················· ····················· ···················· ···················· ····················
G 5283 hCoV-19/Switzerland/SZ-UZH-1000478990/2020|EPI_ISL ···················· ···················· ····················· ···················· ···················· ····················
G 5284 hCoV-19/Switzerland/ZH-UZH-1000479112/2020|EPI_ISL ···················· ···················· ····················· ···················· ···················· ····················
G 5285 hCoV-19/England/BRIS-1337A4/2020|EPI_ISL_524486|20 ···················· ·············A······ ····················· ···················· ···················· ····················
G 5286 hCoV-19/England/BRIS-133883/2020|EPI_ISL_524487|20 ···················· ···················· ·········T··········· ···················· ···················· ····················
G 5287 hCoV-19/England/BRIS-133874/2020|EPI_ISL_524488|20 ···················· ···················· ·········T··········· ···················· ···················· ····················
G 5288 hCoV-19/England/BRIS-13380B/2020|EPI_ISL_524489|20 ···················· ···················· ····················· ···················· ···················· ····················
G 5289 hCoV-19/USA/WA-S2710/2020|EPI_ISL_525700|2020-06-1 ···················· ···················· ····················· ···················· ···················· ····················
G 5290 hCoV-19/USA/WA-S2711/2020|EPI_ISL_525701|2020-06-2 ···················· ···················· ····················· ···················· ···················· ····················
G 5291 hCoV-19/USA/WA-S2712/2020|EPI_ISL_525702|2020-06-2 ···················· ···················· ····················· ···················· ···················· ····················
G 5292 hCoV-19/USA/WA-S2713/2020|EPI_ISL_525703|2020-06-2 ···················· ···················· ····················· ···················· ···················· ····················
G 5293 hCoV-19/USA/WA-S2714/2020|EPI_ISL_525704|2020-06-2 ···················· ···················· ····················· ···················· ···················· ····················
G 5294 hCoV-19/USA/WA-S2715/2020|EPI_ISL_525705|2020-06-2 ···················· ···················· ····················· ···················· ···················· ····················
G 5295 hCoV-19/USA/WA-S2716/2020|EPI_ISL_525706|2020-06-2 ···················· ···················· ····················· ···················· ···················· ····················
G 5296 hCoV-19/USA/WA-S2718/2020|EPI_ISL_525708|2020-07-0 ···················· ···················· ····················· ···················· ···················· ····················
G 5297 hCoV-19/USA/WA-S2719/2020|EPI_ISL_525709|2020-07-0 ···················· ···················· ····················· ···················· ···················· ····················
G 5298 hCoV-19/USA/WA-S2720/2020|EPI_ISL_525710|2020-07-0 ···················· ···················· ····················· ···················· ···················· ····················
G 5299 hCoV-19/USA/WA-S2721/2020|EPI_ISL_525711|2020-07-0 ···················· ···················· ····················· ···················· ···················· ····················
G 5300 hCoV-19/USA/WA-S2722/2020|EPI_ISL_525712|2020-07-0 ···················· ···················· ····················· ···················· ···················· ····················
G 5301 hCoV-19/USA/WA-S2723/2020|EPI_ISL_525713|2020-07-0 ···················· ·A·················· ····················· ···················· ···················· ····················
G 5302 hCoV-19/USA/WA-S2724/2020|EPI_ISL_525714|2020-07-0 ···················· ···················· ····················· ···················· ···················· ····················
G 5303 hCoV-19/USA/WA-S2725/2020|EPI_ISL_525715|2020-07-0 ···················· ···················· ····················· ···················· ···················· ····················
G 5304 hCoV-19/USA/WA-S2726/2020|EPI_ISL_525716|2020-07-0 ···················· ···················· ····················· ···················· ···················· ····················
G 5305 hCoV-19/USA/WA-S2727/2020|EPI_ISL_525717|2020-07-0 ···················· ···················· ····················· ···················· ···················· ····················
G 5306 hCoV-19/USA/WA-S2728/2020|EPI_ISL_525718|2020-07-0 ···················· ···················· ····················· ···················· ···················· ····················
G 5307 hCoV-19/USA/WA-S2729/2020|EPI_ISL_525719|2020-07-0 ···················· ···················· ····················· ···················· ···················· ····················
G 5308 hCoV-19/USA/WA-S2730/2020|EPI_ISL_525720|2020-07-1 ···················· ···················· ····················· ···················· ···················· ····················
G 5309 hCoV-19/USA/WA-S2731/2020|EPI_ISL_525721|2020-07-1 ···················· ···················· ····················· ···················· ···················· ····················
G 5310 hCoV-19/USA/WA-S2732/2020|EPI_ISL_525722|2020-07-1 ···················· ·A····A············· ····················· ···················· ···················· ····················
G 5311 hCoV-19/USA/WA-S2733/2020|EPI_ISL_525723|2020-07-1 ···················· ···················· ····················· ···················· ···················· ····················
G 5312 hCoV-19/USA/WA-S2734/2020|EPI_ISL_525724|2020-07-1 ···················· ···················· ····················· ···················· ···················· ····················
G 5313 hCoV-19/USA/WA-S2735/2020|EPI_ISL_525725|2020-07-1 ···················· ···················· ····················· ···················· ···················· ····················
G 5314 hCoV-19/USA/WA-S2736/2020|EPI_ISL_525726|2020-07-1 ···················· ···················· ····················· ···················· ···················· ····················
G 5315 hCoV-19/USA/WA-S2738/2020|EPI_ISL_525728|2020-07-1 ···················· ···················· ····················· ···················· ···················· ····················
G 5316 hCoV-19/USA/WA-S2739/2020|EPI_ISL_525729|2020-07-1 ···················· ···················· ····················· ···················· ···················· ····················
G 5317 hCoV-19/USA/WA-S2740/2020|EPI_ISL_525730|2020-07-1 ···················· ···················· ····················· ···················· ···················· ····················
G 5318 hCoV-19/USA/WA-S2741/2020|EPI_ISL_525731|2020-07-1 ···················· ···················· ····················· ···················· ···················· ····················
G 5319 hCoV-19/USA/WA-S2742/2020|EPI_ISL_525732|2020-07-1 ···················· ···················· ····················· ···················· ···················· ····················
G 5320 hCoV-19/USA/WA-S2743/2020|EPI_ISL_525733|2020-07-1 ···················· ···················· ····················· ···················· ···················· ····················
G 5321 hCoV-19/USA/WA-S2744/2020|EPI_ISL_525734|2020-07-1 ···················· ···················· ····················· ···················· ···················· ····················
G 5322 hCoV-19/USA/WA-S2745/2020|EPI_ISL_525735|2020-07-1 ···················· ···················· ····················· ···················· ···················· ····················
G 5323 hCoV-19/USA/WA-S2746/2020|EPI_ISL_525736|2020-07-1 ···················· ···················· ····················· ···················· ···················· ····················
G 5324 hCoV-19/USA/WA-S2751/2020|EPI_ISL_525741|2020-07-2 ···················· ···················· ····················· ···················· ···················· ····················
G 5325 hCoV-19/USA/WA-S2752/2020|EPI_ISL_525742|2020-07-2 ···················· ···················· ····················· ···················· ···················· ····················
G 5326 hCoV-19/USA/WA-S2753/2020|EPI_ISL_525743|2020-07-2 ···················· ···················· ····················· ···················· ···················· ····················
G 5327 hCoV-19/USA/WA-S2754/2020|EPI_ISL_525744|2020-07-2 ···················· ···················· ····················· ···················· ···················· ····················
G 5328 hCoV-19/USA/WA-S2756/2020|EPI_ISL_525746|2020-07-2 ···················· ···················· ····················· ···················· ···················· ····················
G 5329 hCoV-19/USA/WA-S2757/2020|EPI_ISL_525747|2020-07-2 ···················· ···················· ····················· ···················· ···················· ····················
G 5330 hCoV-19/USA/WA-S2758/2020|EPI_ISL_525748|2020-07-2 ···················· ···················· ····················· ···················· ···················· ····················
G 5331 hCoV-19/USA/WA-S2759/2020|EPI_ISL_525749|2020-07-2 ···················· ···················· ····················· ···················· ···················· ····················
G 5332 hCoV-19/USA/WA-S2761/2020|EPI_ISL_525751|2020-07-2 ···················· ···················· ····················· ···················· ···················· ····················
G 5333 hCoV-19/USA/WA-S2763/2020|EPI_ISL_525753|2020-07-3 ···················· ···················· ····················· ···················· ···················· ····················
G 5334 hCoV-19/USA/WA-S2764/2020|EPI_ISL_525754|2020-07-3 ···················· ···················· ····················· ···················· ···················· ····················
G 5335 hCoV-19/USA/WA-S2765/2020|EPI_ISL_525755|2020-08-0 ···················· ···················· ····················· ···················· ···················· ····················
G 5336 hCoV-19/USA/WA-S2766/2020|EPI_ISL_525756|2020-07-2 ···················· ···················· ····················· ···················· ···················· ····················
G 5337 hCoV-19/USA/AK-PHL440/2020|EPI_ISL_525758|2020-06- ···················· ···················· ····················· ···················· ···················· ····················

| | | | | | | | | |
|---|---|---|---|---|---|---|---|---|
| G | 5338 | hCoV-19/USA/AK-PHL549/2020\|EPI_ISL_525759\|2020-06- | .................... | .................... | ..................... | .................... | .................... | .................... |
| G | 5339 | hCoV-19/USA/AK-PHL561/2020\|EPI_ISL_525760\|2020-06- | .................... | .................... | ..................... | .................... | .................... | .................... |
| G | 5340 | hCoV-19/USA/TX-DSHS-0200/2020\|EPI_ISL_525763\|2020- | .................... | .................... | ..................... | .................... | .................... | .................... |
| G | 5341 | hCoV-19/USA/TX-DSHS-0237/2020\|EPI_ISL_525788\|2020- | .................... | .................... | ..................... | .................... | .................... | .................... |
| G | 5342 | hCoV-19/Switzerland/BL-UHB-42191545/2020\|EPI_ISL_5 | .................... | .................... | ..................... | .................... | .................... | .................... |
| G | 5343 | hCoV-19/Switzerland/BL-UHB-42191605/2020\|EPI_ISL_5 | .................... | .................... | ..................... | .................... | .................... | .................... |
| G | 5344 | hCoV-19/Switzerland/BL-UHB-42191670/2020\|EPI_ISL_5 | .................... | .................... | ..................... | .................... | .................... | .................... |
| G | 5345 | hCoV-19/Switzerland/BS-UHB-42191689/2020\|EPI_ISL_5 | .................... | .................... | ..................... | .................... | .................... | .................... |
| G | 5346 | hCoV-19/Switzerland/BL-UHB-42191693/2020\|EPI_ISL_5 | .................... | .................... | ..................... | .................... | .................... | .................... |
| G | 5347 | hCoV-19/Switzerland/BL-UHB-42191735/2020\|EPI_ISL_5 | .................... | .................... | ..................... | .................... | .................... | .................... |
| G | 5348 | hCoV-19/Switzerland/BL-UHB-42191739/2020\|EPI_ISL_5 | .................... | .................... | ..................... | .................... | .................... | .................... |
| G | 5349 | hCoV-19/Switzerland/BL-UHB-42191742/2020\|EPI_ISL_5 | .................... | .................... | ..................... | .................... | .................... | .................... |
| G | 5350 | hCoV-19/Switzerland/BS-UHB-42191744/2020\|EPI_ISL_5 | .................... | .................... | ..................... | .................... | .NN................. | .................... |
| G | 5351 | hCoV-19/Switzerland/BS-UHB-42191751/2020\|EPI_ISL_5 | .................... | .................... | ..................... | .................... | .................... | .................... |
| G | 5352 | hCoV-19/Switzerland/BL-UHB-42191804/2020\|EPI_ISL_5 | .................... | .................... | ..................... | .................... | .................... | .................... |
| G | 5353 | hCoV-19/Switzerland/BS-UHB-42191841/2020\|EPI_ISL_5 | .................... | .................... | ..................... | .................... | .................... | .................... |
| G | 5354 | hCoV-19/Switzerland/BS-UHB-42191874/2020\|EPI_ISL_5 | .................... | .................... | ..................... | .................... | .................... | .................... |
| G | 5355 | hCoV-19/Switzerland/BL-UHB-42191950/2020\|EPI_ISL_5 | .................... | .................... | ..................... | .................... | .................... | .................... |
| G | 5356 | hCoV-19/Switzerland/BS-UHB-42191965/2020\|EPI_ISL_5 | .................... | .................... | ..................... | .................... | .................... | .................... |
| G | 5357 | hCoV-19/Switzerland/BS-UHB-42191987/2020\|EPI_ISL_5 | .................... | .................... | ..................... | .................... | .................... | .................... |
| G | 5358 | hCoV-19/Switzerland/BS-UHB-42192008/2020\|EPI_ISL_5 | .................... | .................... | ..................... | .................... | .................... | .................... |
| G | 5359 | hCoV-19/Switzerland/BS-UHB-42192010/2020\|EPI_ISL_5 | .................... | .................... | ..................... | .................... | .................... | .................... |
| G | 5360 | hCoV-19/Switzerland/BL-UHB-42192025/2020\|EPI_ISL_5 | .................... | .................... | ..................... | .................... | .................... | .................... |
| G | 5361 | hCoV-19/Switzerland/BS-UHB-42192039/2020\|EPI_ISL_5 | .................... | .................... | ..................... | .................... | .................... | .................... |
| G | 5362 | hCoV-19/Switzerland/BS-UHB-42192047/2020\|EPI_ISL_5 | .................... | .................... | ..................... | .................... | .................... | .................... |
| G | 5363 | hCoV-19/Switzerland/BL-UHB-42192108/2020\|EPI_ISL_5 | .................... | .................... | ..................... | .................... | .................... | .................... |
| G | 5364 | hCoV-19/Switzerland/BL-UHB-42192277/2020\|EPI_ISL_5 | .................... | .................... | ..................... | .................... | .................... | .................... |
| G | 5365 | hCoV-19/Switzerland/BS-UHB-42192489/2020\|EPI_ISL_5 | .................... | .................... | ..................... | .................... | .................... | .................... |
| G | 5366 | hCoV-19/Switzerland/BS-UHB-42192492/2020\|EPI_ISL_5 | .................... | .................... | ..................... | .................... | .................... | .................... |
| G | 5367 | hCoV-19/Switzerland/BS-UHB-42192764/2020\|EPI_ISL_5 | .................... | .................... | ..................... | .................... | .................... | .................... |
| G | 5368 | hCoV-19/Switzerland/BL-UHB-42192815/2020\|EPI_ISL_5 | .................... | .................... | ..................... | .................... | .................... | .................... |
| G | 5369 | hCoV-19/Switzerland/BL-UHB-42192855/2020\|EPI_ISL_5 | .................... | .................... | ..................... | .................... | .................... | .................... |
| G | 5370 | hCoV-19/Switzerland/BS-UHB-42192872/2020\|EPI_ISL_5 | .................... | .................... | ..................... | .................... | .................... | .................... |
| G | 5371 | hCoV-19/Switzerland/BL-UHB-42192884/2020\|EPI_ISL_5 | .................... | .................... | ..................... | .................... | .................... | .................... |
| G | 5372 | hCoV-19/Switzerland/BS-UHB-42192894/2020\|EPI_ISL_5 | .................... | .................... | ..................... | .................... | .................... | .................... |
| G | 5373 | hCoV-19/Switzerland/BL-UHB-42192918/2020\|EPI_ISL_5 | .................... | .................... | ..................... | .................... | .................... | .................... |
| G | 5374 | hCoV-19/Switzerland/BL-UHB-42192943/2020\|EPI_ISL_5 | .................... | .................... | ..................... | .................... | .................... | .................... |
| G | 5375 | hCoV-19/Switzerland/BS-UHB-42192965/2020\|EPI_ISL_5 | .................... | .................... | ..................... | .................... | .................... | .................... |
| G | 5376 | hCoV-19/Switzerland/BL-UHB-42192973/2020\|EPI_ISL_5 | .................... | .................... | ..................... | .................... | .................... | .................... |
| G | 5378 | hCoV-19/England/LIVE-A99A4/2020\|EPI_ISL_517301\|202 | .................... | .................... | ..................... | .................... | .................... | .................... |
| G | 5379 | hCoV-19/England/LIVE-A99B3/2020\|EPI_ISL_517302\|202 | .................... | .................... | ..................... | .................... | .................... | .................... |
| G | 5380 | hCoV-19/England/LIVE-A99C2/2020\|EPI_ISL_517303\|202 | .................... | .................... | ..................... | .................... | .................... | .................... |
| G | 5382 | hCoV-19/England/LIVE-A99E0/2020\|EPI_ISL_517305\|202 | .................... | .................... | ..................... | .................... | .................... | .................... |
| G | 5384 | hCoV-19/England/LIVE-A9A0B/2020\|EPI_ISL_517307\|202 | .................... | .................... | ..................... | .................... | ......NN............ | .................... |
| G | 5385 | hCoV-19/England/LIVE-A9A1A/2020\|EPI_ISL_517308\|202 | .................... | .................... | ..................... | .................... | .................... | .................... |
| G | 5386 | hCoV-19/England/LIVE-A9A29/2020\|EPI_ISL_517309\|202 | .................... | .................... | ..................... | .................... | .................... | .................... |
| G | 5387 | hCoV-19/England/LIVE-A9A38/2020\|EPI_ISL_517310\|202 | .................... | .................... | ..................... | .................... | .................... | .................... |
| G | 5388 | hCoV-19/England/LIVE-A9A47/2020\|EPI_ISL_517311\|202 | .................... | .................... | ..................... | .................... | .................... | .................... |
| G | 5389 | hCoV-19/England/LIVE-A9A56/2020\|EPI_ISL_517312\|202 | .................... | .................... | ..................... | .................... | .................... | .................... |
| G | 5390 | hCoV-19/England/LIVE-A9A65/2020\|EPI_ISL_517313\|202 | .................... | .................... | ..................... | .................... | .................... | .................... |
| G | 5391 | hCoV-19/England/LIVE-A9A74/2020\|EPI_ISL_517314\|202 | .................... | .................... | ..................... | .................... | .................... | .................... |
| G | 5392 | hCoV-19/England/LIVE-A9A83/2020\|EPI_ISL_517315\|202 | .................... | .................... | ..................... | .................... | .................... | .................... |
| G | 5395 | hCoV-19/England/LIVE-A9AB0/2020\|EPI_ISL_517318\|202 | .................... | .................... | ..................... | .................... | .................... | .................... |
| G | 5396 | hCoV-19/England/LIVE-A9ACF/2020\|EPI_ISL_517319\|202 | .................... | .................... | ..................... | .................... | .................... | .................... |
| G | 5397 | hCoV-19/Australia/VIC5805/2020\|EPI_ISL_518600\|2020 | .................... | .................... | ..................... | .................... | .................... | .................... |
| G | 5398 | hCoV-19/Australia/VIC5806/2020\|EPI_ISL_518601\|2020 | .................... | .................... | ..................... | .................... | .................... | .................... |
| G | 5399 | hCoV-19/Australia/VIC5807/2020\|EPI_ISL_518602\|2020 | .................... | .................... | ..................... | .................... | .................... | .................... |
| G | 5400 | hCoV-19/Australia/VIC5808/2020\|EPI_ISL_518603\|2020 | .................... | .................... | ..................... | .................... | .................... | .................... |
| G | 5401 | hCoV-19/Australia/VIC5809/2020\|EPI_ISL_518604\|2020 | .................... | .................... | ..................... | .................... | .................... | .................... |
| G | 5402 | hCoV-19/Australia/VIC5810/2020\|EPI_ISL_518605\|2020 | .................... | .................... | ..................... | .................... | .................... | .................... |
| G | 5403 | hCoV-19/Australia/VIC5811/2020\|EPI_ISL_518606\|2020 | .................... | .................... | ..................... | .................... | .................... | .................... |
| G | 5404 | hCoV-19/Australia/VIC5812/2020\|EPI_ISL_518607\|2020 | .................... | .................... | ..................... | .................... | .................... | .................... |
| G | 5405 | hCoV-19/Australia/VIC5813/2020\|EPI_ISL_518608\|2020 | .................... | .................... | ..................... | .................... | .................... | .................... |
| G | 5406 | hCoV-19/Australia/VIC5814/2020\|EPI_ISL_518609\|2020 | .................... | .................... | ..................... | .................... | .................... | .................... |
| G | 5407 | hCoV-19/Australia/VIC5816/2020\|EPI_ISL_518610\|2020 | .................... | .................... | ..................... | .................... | .................... | .................... |
| G | 5408 | hCoV-19/Australia/VIC5817/2020\|EPI_ISL_518611\|2020 | .................... | .................... | ..................... | .................... | .................... | .................... |
| G | 5409 | hCoV-19/Australia/VIC5818/2020\|EPI_ISL_518612\|2020 | .................... | .................... | ..................... | .................... | .................... | .................... |
| G | 5410 | hCoV-19/Australia/VIC5819/2020\|EPI_ISL_518613\|2020 | .................... | .................... | ..................... | .................... | .................... | .................... |
| G | 5411 | hCoV-19/Australia/VIC5820/2020\|EPI_ISL_518614\|2020 | .................... | .................... | ..................... | .................... | .................... | .................... |
| G | 5412 | hCoV-19/Australia/VIC5821/2020\|EPI_ISL_518615\|2020 | .................... | .................... | ..................... | .................... | .................... | .................... |
| G | 5413 | hCoV-19/Australia/VIC5822/2020\|EPI_ISL_518616\|2020 | .................... | .................... | ..................... | .................... | .................... | .................... |
| G | 5414 | hCoV-19/Australia/VIC5823/2020\|EPI_ISL_518617\|2020 | .................... | .................... | ..................... | .................... | .................... | ...A................ |
| G | 5415 | hCoV-19/Australia/VIC5824/2020\|EPI_ISL_518618\|2020 | .................... | .................... | ..................... | .................... | .................... | .................... |
| G | 5416 | hCoV-19/Australia/VIC5825/2020\|EPI_ISL_518619\|2020 | .................... | .................... | ..................... | .................... | .................... | .................... |
| G | 5417 | hCoV-19/Australia/VIC5826/2020\|EPI_ISL_518620\|2020 | .................... | .................... | ..................... | .................... | .................... | .................... |
| G | 5418 | hCoV-19/Australia/VIC5827/2020\|EPI_ISL_518621\|2020 | .................... | .................... | ..................... | .................... | .................... | .................... |
| G | 5419 | hCoV-19/Australia/VIC5828/2020\|EPI_ISL_518622\|2020 | .................... | .................... | ..................... | .................... | .................... | .................... |
| G | 5420 | hCoV-19/Australia/VIC5829/2020\|EPI_ISL_518623\|2020 | .................... | .................... | ..................... | .................... | .................... | .................... |
| G | 5421 | hCoV-19/Australia/VIC5830/2020\|EPI_ISL_518624\|2020 | .................... | .................... | ..................... | .................... | .................... | .................... |
| G | 5422 | hCoV-19/Australia/VIC5831/2020\|EPI_ISL_518625\|2020 | .................... | .................... | ..................... | .................... | .................... | .................... |

| | | | | | | | |
|---|---|---|---|---|---|---|---|
| G | 5423 hCoV-19/Australia/VIC5832/2020\|EPI_ISL_518626\|2020 | .................... | .................... | ..................... | .................... | .................... | .................... |
| G | 5424 hCoV-19/Australia/VIC5833/2020\|EPI_ISL_518627\|2020 | .................... | .................... | ..................... | .................... | .................... | .................... |
| G | 5425 hCoV-19/Australia/VIC5834/2020\|EPI_ISL_518628\|2020 | .................... | .................... | ..................... | .................... | .................... | .................... |
| G | 5426 hCoV-19/Australia/VIC5835/2020\|EPI_ISL_518629\|2020 | .................... | .................... | ..................... | .................... | .................... | .................... |
| G | 5427 hCoV-19/Australia/VIC5836/2020\|EPI_ISL_518630\|2020 | .................... | .................... | ..................... | .................... | .................... | .................... |
| G | 5428 hCoV-19/Australia/VIC5837/2020\|EPI_ISL_518631\|2020 | .................... | .................... | ..................... | .................... | .................... | .................... |
| G | 5429 hCoV-19/Australia/VIC5838/2020\|EPI_ISL_518632\|2020 | .................... | .................... | ..................... | .................... | .................... | .................... |
| G | 5430 hCoV-19/Australia/VIC5839/2020\|EPI_ISL_518633\|2020 | .................... | .................... | ..................... | .................... | .................... | .................... |
| G | 5431 hCoV-19/Australia/VIC5840/2020\|EPI_ISL_518634\|2020 | .................... | .................... | ..................... | .................... | .................... | .................... |
| G | 5432 hCoV-19/Australia/VIC5841/2020\|EPI_ISL_518635\|2020 | .................... | .................... | ..................... | .................... | .................... | .................... |
| G | 5433 hCoV-19/Australia/VIC5842/2020\|EPI_ISL_518636\|2020 | .................... | .................... | ..................... | .................... | .................... | .................... |
| G | 5434 hCoV-19/Australia/VIC5843/2020\|EPI_ISL_518637\|2020 | .................... | .................... | ..................... | .................... | .................... | .................... |
| G | 5435 hCoV-19/Australia/VIC5844/2020\|EPI_ISL_518638\|2020 | .................... | .................... | ..................... | .................... | .................... | .................... |
| G | 5436 hCoV-19/Australia/VIC5845/2020\|EPI_ISL_518639\|2020 | .................... | .................... | ..................... | .................... | .................... | .................... |
| G | 5437 hCoV-19/Australia/VIC5846/2020\|EPI_ISL_518640\|2020 | .................... | .................... | ..................... | .................... | .................... | .................... |
| G | 5438 hCoV-19/Australia/VIC5847/2020\|EPI_ISL_518641\|2020 | .................... | .................... | ..................... | .................... | .................... | .................... |
| G | 5439 hCoV-19/Australia/VIC5849/2020\|EPI_ISL_518642\|2020 | .................... | .................... | ..................... | .................... | .................... | .................... |
| G | 5440 hCoV-19/Australia/VIC5850/2020\|EPI_ISL_518643\|2020 | .................... | .................... | ..................... | .................... | .................... | .................... |
| G | 5441 hCoV-19/Australia/VIC5851/2020\|EPI_ISL_518644\|2020 | .................... | .................... | ..................... | .................... | .................... | .................... |
| G | 5443 hCoV-19/Australia/VIC5853/2020\|EPI_ISL_518646\|2020 | .................... | .................... | ..................... | .................... | .................... | .................... |
| G | 5444 hCoV-19/Australia/VIC5854/2020\|EPI_ISL_518647\|2020 | .................... | .................... | ..................... | .................... | .................... | .................... |
| G | 5445 hCoV-19/Australia/VIC5856/2020\|EPI_ISL_518649\|2020 | .................... | .................... | ..................... | .................... | .................... | .................... |
| G | 5446 hCoV-19/Australia/VIC5097/2020\|EPI_ISL_519900\|2020 | .................... | .................... | ..................... | .................... | .................... | .................... |
| G | 5447 hCoV-19/Australia/VIC5098/2020\|EPI_ISL_519901\|2020 | .................... | .................... | ..................... | .................... | .................... | .................... |
| G | 5448 hCoV-19/Australia/VIC5099/2020\|EPI_ISL_519902\|2020 | .................... | .................... | ..................... | .................... | .................... | .................... |
| G | 5449 hCoV-19/Australia/VIC5100/2020\|EPI_ISL_519903\|2020 | .................... | .................... | ..................... | .................... | .................... | .................... |
| G | 5450 hCoV-19/Australia/VIC5101/2020\|EPI_ISL_519904\|2020 | .................... | .................... | ..................... | .................... | .................... | .................... |
| G | 5451 hCoV-19/Australia/VIC5102/2020\|EPI_ISL_519905\|2020 | .................... | .................... | ..................... | .................... | .................... | .................... |
| G | 5452 hCoV-19/Australia/VIC5103/2020\|EPI_ISL_519906\|2020 | .................... | .................... | ..................... | .................... | .................... | .................... |
| G | 5453 hCoV-19/Australia/VIC5104/2020\|EPI_ISL_519907\|2020 | .................... | .................... | ..................... | .................... | .................... | .................... |
| G | 5454 hCoV-19/Australia/VIC5105/2020\|EPI_ISL_519908\|2020 | .................... | .................... | ..................... | .................... | .................... | .................... |
| G | 5455 hCoV-19/Australia/VIC5106/2020\|EPI_ISL_519909\|2020 | .................... | .................... | ..................... | .................... | .................... | .................... |
| G | 5456 hCoV-19/Australia/VIC5107/2020\|EPI_ISL_519910\|2020 | .................... | .................... | ..................... | .................... | .................... | .................... |
| G | 5457 hCoV-19/Australia/VIC5108/2020\|EPI_ISL_519911\|2020 | .................... | .................... | ..................... | .................... | .................... | .................... |
| G | 5458 hCoV-19/Australia/VIC5109/2020\|EPI_ISL_519912\|2020 | .................... | .................... | ..................... | .................... | .................... | .................... |
| G | 5459 hCoV-19/Australia/VIC5110/2020\|EPI_ISL_519913\|2020 | .................... | .................... | ..................... | .................... | .................... | .................... |
| G | 5460 hCoV-19/Australia/VIC5111/2020\|EPI_ISL_519914\|2020 | .................... | .................... | ..................... | .................... | .................... | .................... |
| G | 5461 hCoV-19/Australia/VIC5112/2020\|EPI_ISL_519915\|2020 | .................... | .................... | ..................... | .................... | .................... | .................... |
| G | 5462 hCoV-19/Australia/VIC5113/2020\|EPI_ISL_519916\|2020 | .................... | .................... | ..................... | .................... | .................... | .................... |
| G | 5463 hCoV-19/Australia/VIC5114/2020\|EPI_ISL_519917\|2020 | .................... | .................... | ..................... | .................... | .................... | .................... |
| G | 5464 hCoV-19/Australia/VIC5115/2020\|EPI_ISL_519918\|2020 | .................... | .................... | ..................... | .................... | .................... | .................... |
| G | 5465 hCoV-19/Australia/VIC5116/2020\|EPI_ISL_519919\|2020 | .................... | .................... | ..................... | .................... | .................... | .................... |
| G | 5466 hCoV-19/Australia/VIC5117/2020\|EPI_ISL_519920\|2020 | .................... | .................... | ..................... | .................... | .................... | .................... |
| G | 5467 hCoV-19/Australia/VIC5118/2020\|EPI_ISL_519921\|2020 | .................... | .................... | ..................... | .................... | .................... | .................... |
| G | 5468 hCoV-19/Australia/VIC5119/2020\|EPI_ISL_519922\|2020 | .................... | .................... | ..................... | .................... | .................... | .................... |
| G | 5469 hCoV-19/Australia/VIC5120/2020\|EPI_ISL_519923\|2020 | .................... | .................... | ..................... | .................... | .................... | .................... |
| G | 5470 hCoV-19/Australia/VIC5121/2020\|EPI_ISL_519924\|2020 | .................... | .................... | ..................... | .................... | .................... | .................... |
| G | 5471 hCoV-19/Australia/VIC5123/2020\|EPI_ISL_519926\|2020 | .................... | .................... | ..................... | .................... | .................... | .................... |
| G | 5472 hCoV-19/Australia/VIC5124/2020\|EPI_ISL_519927\|2020 | .................... | .................... | ..................... | .................... | .................... | .................... |
| G | 5473 hCoV-19/Australia/VIC5125/2020\|EPI_ISL_519928\|2020 | .................... | .................... | ..................... | .................... | .................... | .................... |
| G | 5474 hCoV-19/Australia/VIC5126/2020\|EPI_ISL_519929\|2020 | .................... | .................... | ..................... | .................... | .................... | .................... |
| G | 5475 hCoV-19/Australia/VIC5127/2020\|EPI_ISL_519930\|2020 | .................... | .................... | ..................... | .................... | .................... | .................... |
| G | 5476 hCoV-19/Australia/VIC5128/2020\|EPI_ISL_519931\|2020 | .................... | .................... | ..................... | .................... | .................... | .................... |
| G | 5477 hCoV-19/Australia/VIC5129/2020\|EPI_ISL_519932\|2020 | .................... | .................... | ..................... | .................... | .................... | .................... |
| G | 5478 hCoV-19/Australia/VIC5130/2020\|EPI_ISL_519933\|2020 | .................... | .................... | ..................... | .................... | .................... | .................... |
| G | 5479 hCoV-19/Australia/VIC5131/2020\|EPI_ISL_519934\|2020 | .................... | .................... | ..................... | .................... | .................... | .................... |
| G | 5480 hCoV-19/Australia/VIC5132/2020\|EPI_ISL_519935\|2020 | .................... | .................... | ..................... | .................... | .................... | .................... |
| G | 5481 hCoV-19/Australia/VIC5133/2020\|EPI_ISL_519936\|2020 | .................... | .................... | ..................... | .................... | .................... | .................... |
| G | 5482 hCoV-19/Australia/VIC5134/2020\|EPI_ISL_519937\|2020 | .................... | .................... | ..................... | .................... | .................... | .................... |
| G | 5483 hCoV-19/Australia/VIC5135/2020\|EPI_ISL_519938\|2020 | .................... | .................... | ..................... | .................... | .................... | .................... |
| G | 5484 hCoV-19/Australia/VIC5136/2020\|EPI_ISL_519939\|2020 | .................... | .................... | ..................... | .................... | .................... | .................... |
| G | 5485 hCoV-19/Australia/VIC5137/2020\|EPI_ISL_519940\|2020 | .................... | .................... | ..................... | .................... | .................... | .................... |
| G | 5486 hCoV-19/Australia/VIC5138/2020\|EPI_ISL_519941\|2020 | .................... | .................... | ..................... | .................... | .................... | .................... |
| G | 5487 hCoV-19/Australia/VIC5139/2020\|EPI_ISL_519942\|2020 | .................... | .................... | ..................... | .................... | .................... | .................... |
| G | 5488 hCoV-19/Australia/VIC5140/2020\|EPI_ISL_519943\|2020 | .................... | .................... | ..................... | .................... | .................... | .................... |
| G | 5489 hCoV-19/Australia/VIC5141/2020\|EPI_ISL_519944\|2020 | .................... | .................... | ..................... | .................... | .................... | .................... |
| G | 5490 hCoV-19/Australia/VIC5142/2020\|EPI_ISL_519945\|2020 | .................... | .................... | ..................... | .................... | .................... | .................... |
| G | 5491 hCoV-19/Australia/VIC5143/2020\|EPI_ISL_519946\|2020 | .................... | .................... | ..................... | .................... | .................... | .................... |
| G | 5492 hCoV-19/Australia/VIC5145/2020\|EPI_ISL_519947\|2020 | .................... | .................... | ..................... | .................... | .................... | .................... |
| G | 5493 hCoV-19/Australia/VIC5150/2020\|EPI_ISL_519948\|2020 | .................... | .................... | ..................... | .................... | .................... | .................... |
| G | 5494 hCoV-19/Australia/VIC5180/2020\|EPI_ISL_519950\|2020 | .................... | .................... | ..................... | .................... | .................... | .................... |
| G | 5495 hCoV-19/Australia/VIC5186/2020\|EPI_ISL_519951\|2020 | .................... | .................... | ..................... | .................... | .................... | .................... |
| G | 5496 hCoV-19/Australia/VIC5187/2020\|EPI_ISL_519952\|2020 | .................... | .................... | ..................... | .................... | .................... | .................... |
| G | 5497 hCoV-19/Australia/VIC5188/2020\|EPI_ISL_519953\|2020 | .................... | .................... | ..................... | .................... | .................... | .................... |
| G | 5498 hCoV-19/Australia/VIC5189/2020\|EPI_ISL_519954\|2020 | .................... | .................... | ..................... | .................... | .................... | .................... |
| G | 5499 hCoV-19/Australia/VIC5190/2020\|EPI_ISL_519955\|2020 | .................... | .................... | ..................... | .................... | .................... | .................... |
| G | 5500 hCoV-19/Australia/VIC5191/2020\|EPI_ISL_519956\|2020 | .................... | .................... | ..................... | .................... | .................... | .................... |
| G | 5501 hCoV-19/Australia/VIC5192/2020\|EPI_ISL_519957\|2020 | .................... | .................... | ..................... | .................... | .................... | .................... |
| G | 5502 hCoV-19/Australia/VIC5193/2020\|EPI_ISL_519958\|2020 | .................... | .................... | ..................... | .................... | .................... | .................... |
| G | 5503 hCoV-19/Australia/VIC5194/2020\|EPI_ISL_519959\|2020 | .................... | .................... | ..................... | .................... | .................... | .................... |

| | | | | | | | | |
|---|---|---|---|---|---|---|---|---|
| G | 5504 hCoV-19/Australia/VIC5195/2020\|EPI_ISL_519960\|2020 | ···················· | ···················· | ···················· | ···················· | ···················· | ···················· | |
| G | 5505 hCoV-19/Australia/VIC5196/2020\|EPI_ISL_519961\|2020 | ···················· | ···················· | ···················· | ···················· | ···················· | ···················· | |
| G | 5506 hCoV-19/Australia/VIC5197/2020\|EPI_ISL_519962\|2020 | ···················· | ···················· | ···················· | ···················· | ···················· | ···················· | |
| G | 5507 hCoV-19/Australia/VIC5198/2020\|EPI_ISL_519963\|2020 | ···················· | ···················· | ···················· | ···················· | ···················· | ···················· | |
| G | 5508 hCoV-19/Australia/VIC5199/2020\|EPI_ISL_519964\|2020 | ···················· | ···················· | ···················· | ···················· | ···················· | ···················· | |
| G | 5509 hCoV-19/Australia/VIC5200/2020\|EPI_ISL_519965\|2020 | ···················· | ···················· | ···················· | ···················· | ···················· | ···················· | |
| G | 5510 hCoV-19/Australia/VIC5201/2020\|EPI_ISL_519966\|2020 | ···················· | ···················· | ···················· | ···················· | ···················· | ···················· | |
| G | 5511 hCoV-19/Australia/VIC5202/2020\|EPI_ISL_519967\|2020 | ···················· | ···················· | ···················· | ···················· | ···················· | ···················· | |
| G | 5512 hCoV-19/Australia/VIC5203/2020\|EPI_ISL_519968\|2020 | ···················· | ···················· | ···················· | ···················· | ···················· | ···················· | |
| G | 5513 hCoV-19/Australia/VIC5204/2020\|EPI_ISL_519969\|2020 | ···················· | ···················· | ···················· | ···················· | ···················· | ···················· | |
| G | 5514 hCoV-19/Australia/VIC5206/2020\|EPI_ISL_519971\|2020 | ···················· | ···················· | ···················· | ···················· | ···················· | ···················· | |
| G | 5515 hCoV-19/Australia/VIC5207/2020\|EPI_ISL_519972\|2020 | ···················· | ···················· | ···················· | ···················· | ···················· | ···················· | |
| G | 5516 hCoV-19/Australia/VIC5208/2020\|EPI_ISL_519973\|2020 | ···················· | ···················· | ···················· | ···················· | ···················· | ···················· | |
| G | 5517 hCoV-19/Australia/VIC5209/2020\|EPI_ISL_519974\|2020 | ···················· | ···················· | ···················· | ···················· | ···················· | ···················· | |
| G | 5518 hCoV-19/Australia/VIC5210/2020\|EPI_ISL_519975\|2020 | ···················· | ···················· | ···················· | ···················· | ···················· | ···················· | |
| G | 5519 hCoV-19/Australia/VIC5211/2020\|EPI_ISL_519976\|2020 | ···················· | ···················· | ···················· | ···················· | ···················· | ···················· | |
| G | 5520 hCoV-19/Australia/VIC5212/2020\|EPI_ISL_519977\|2020 | ···················· | ···················· | ···················· | ···················· | ···················· | ···················· | |
| G | 5521 hCoV-19/Australia/VIC5213/2020\|EPI_ISL_519978\|2020 | ···················· | ···················· | ···················· | ···················· | ···················· | ···················· | |
| G | 5522 hCoV-19/Australia/VIC5214/2020\|EPI_ISL_519979\|2020 | ···················· | ···················· | ···················· | ···················· | ···················· | ···················· | |
| G | 5523 hCoV-19/Australia/VIC3215/2020\|EPI_ISL_520900\|2020 | ···················· | ···················· | ···················· | ···················· | ···················· | ···················· | |
| G | 5524 hCoV-19/Australia/VIC3216/2020\|EPI_ISL_520901\|2020 | ···················· | ···················· | ···················· | ···················· | ···················· | ···················· | |
| G | 5525 hCoV-19/Australia/VIC3217/2020\|EPI_ISL_520902\|2020 | ···················· | ···················· | ···················· | ···················· | ···················· | ···················· | |
| G | 5527 hCoV-19/Australia/VIC3219/2020\|EPI_ISL_520904\|2020 | ···················· | ···················· | ···················· | ···················· | ···················· | ···················· | |
| G | 5528 hCoV-19/Australia/VIC3220/2020\|EPI_ISL_520905\|2020 | ···················· | ···················· | ···················· | ···················· | ···················· | ···················· | |
| G | 5529 hCoV-19/Australia/VIC3223/2020\|EPI_ISL_520906\|2020 | ···················· | ···················· | ···················· | ···················· | ···················· | ···················· | |
| G | 5530 hCoV-19/Australia/VIC3224/2020\|EPI_ISL_520907\|2020 | ···················· | ···················· | ···················· | ···················· | ···················· | ···················· | |
| G | 5531 hCoV-19/Australia/VIC3225/2020\|EPI_ISL_520908\|2020 | ···················· | ···················· | ···················· | ···················· | ···················· | ···················· | |
| G | 5532 hCoV-19/Australia/VIC3226/2020\|EPI_ISL_520909\|2020 | ···················· | ···················· | ···················· | ···················· | ···················· | ···················· | |
| G | 5533 hCoV-19/Australia/VIC3227/2020\|EPI_ISL_520910\|2020 | ···················· | ···················· | ···················· | ···················· | ···················· | ···················· | |
| G | 5534 hCoV-19/Australia/VIC3228/2020\|EPI_ISL_520911\|2020 | ···················· | ···················· | ···················· | ···················· | ···················· | ···················· | |
| G | 5535 hCoV-19/Australia/VIC3230/2020\|EPI_ISL_520913\|2020 | ···················· | ···················· | ···················· | ···················· | ···················· | ···················· | |
| G | 5536 hCoV-19/Australia/VIC3231/2020\|EPI_ISL_520914\|2020 | ···················· | ···················· | ···················· | ···················· | ·NNN················ | ···················· | 1 |
| G | 5537 hCoV-19/Australia/VIC3233/2020\|EPI_ISL_520915\|2020 | ···················· | ···················· | ···················· | ···················· | ···················· | ···················· | |
| G | 5538 hCoV-19/Australia/VIC3235/2020\|EPI_ISL_520916\|2020 | ···················· | ···················· | ···················· | ···················· | ···················· | ···················· | |
| G | 5539 hCoV-19/Australia/VIC3236/2020\|EPI_ISL_520917\|2020 | ···················· | ···················· | ···················· | ···················· | ···················· | ···················· | |
| G | 5540 hCoV-19/Australia/VIC3237/2020\|EPI_ISL_520918\|2020 | ···················· | ···················· | ···················· | ···················· | ···················· | ···················· | |
| G | 5541 hCoV-19/Australia/VIC3238/2020\|EPI_ISL_520919\|2020 | ···················· | ···················· | ···················· | ···················· | ···················· | ···················· | |
| G | 5542 hCoV-19/Australia/VIC3239/2020\|EPI_ISL_520920\|2020 | ···················· | ···················· | ···················· | ···················· | ···················· | ···················· | |
| G | 5543 hCoV-19/Australia/VIC3240/2020\|EPI_ISL_520921\|2020 | ···················· | ···················· | ···················· | ···················· | ···················· | ···················· | |
| G | 5544 hCoV-19/Australia/VIC3241/2020\|EPI_ISL_520922\|2020 | ···················· | ···················· | ···················· | ···················· | ···················· | ···················· | |
| G | 5545 hCoV-19/Australia/VIC3242/2020\|EPI_ISL_520923\|2020 | ···················· | ···················· | ···················· | ···················· | ···················· | ···················· | |
| G | 5546 hCoV-19/Australia/VIC3243/2020\|EPI_ISL_520924\|2020 | ···················· | ·········A·········· | ···················· | ···················· | ···················· | ···················· | |
| G | 5547 hCoV-19/Australia/VIC3245/2020\|EPI_ISL_520925\|2020 | ···················· | ···················· | ···················· | ···················· | ···················· | ···················· | |
| G | 5548 hCoV-19/Australia/VIC3246/2020\|EPI_ISL_520926\|2020 | ···················· | ···················· | ···················· | ···················· | ···················· | ···················· | |
| G | 5549 hCoV-19/Australia/VIC3247/2020\|EPI_ISL_520927\|2020 | ···················· | ···················· | ···················· | ···················· | ···················· | ···················· | |
| G | 5550 hCoV-19/Australia/VIC3248/2020\|EPI_ISL_520928\|2020 | ···················· | ···················· | ···················· | ···················· | ···················· | ···················· | |
| G | 5551 hCoV-19/Australia/VIC3249/2020\|EPI_ISL_520929\|2020 | ···················· | ···················· | ···················· | ···················· | ···················· | ···················· | |
| G | 5552 hCoV-19/Australia/VIC3250/2020\|EPI_ISL_520930\|2020 | ···················· | ···················· | ···················· | ···················· | ···················· | ···················· | |
| G | 5553 hCoV-19/Australia/VIC3251/2020\|EPI_ISL_520931\|2020 | ···················· | ···················· | ···················· | ···················· | ···················· | ···················· | |
| G | 5554 hCoV-19/Australia/VIC3252/2020\|EPI_ISL_520932\|2020 | ···················· | ···················· | ···················· | ···················· | ···················· | ···················· | |
| G | 5555 hCoV-19/Australia/VIC3253/2020\|EPI_ISL_520933\|2020 | ···················· | ···················· | ···················· | ···················· | NNN················· | ···················· | 1 |
| G | 5557 hCoV-19/Australia/VIC3255/2020\|EPI_ISL_520935\|2020 | ···················· | ···················· | ···················· | ···················· | ···················· | ···················· | |
| G | 5558 hCoV-19/Australia/VIC3256/2020\|EPI_ISL_520936\|2020 | ···················· | ···················· | ···················· | ···················· | ···················· | ···················· | |
| G | 5560 hCoV-19/Australia/VIC3259/2020\|EPI_ISL_520938\|2020 | ···················· | ···················· | ···················· | ···················· | ···················· | ···················· | |
| G | 5561 hCoV-19/Australia/VIC3260/2020\|EPI_ISL_520939\|2020 | ···················· | ···················· | ···················· | ···················· | ···················· | ···················· | |
| G | 5562 hCoV-19/Australia/VIC3261/2020\|EPI_ISL_520940\|2020 | ···················· | ···················· | ···················· | ···················· | ···················· | ···················· | |
| G | 5563 hCoV-19/Australia/VIC3262/2020\|EPI_ISL_520941\|2020 | ···················· | ···················· | ···················· | ···················· | ···················· | ···················· | |
| G | 5566 hCoV-19/Australia/VIC3265/2020\|EPI_ISL_520944\|2020 | ···················· | ···················· | ···················· | ···················· | ···················· | ···················· | |
| G | 5567 hCoV-19/Australia/VIC3266/2020\|EPI_ISL_520945\|2020 | ···················· | ···················· | ···················· | ···················· | ···················· | ···················· | |
| G | 5568 hCoV-19/Australia/VIC3267/2020\|EPI_ISL_520946\|2020 | ···················· | ···················· | ···················· | ···················· | ···················· | ···················· | |
| G | 5569 hCoV-19/Australia/VIC3268/2020\|EPI_ISL_520947\|2020 | ···················· | ···················· | ···················· | ···················· | ···················· | ···················· | |
| G | 5570 hCoV-19/Australia/VIC3270/2020\|EPI_ISL_520949\|2020 | ···················· | ···················· | ···················· | ···················· | ···················· | ···················· | |
| G | 5571 hCoV-19/Australia/VIC3271/2020\|EPI_ISL_520950\|2020 | ···················· | ···················· | ···················· | ···················· | ···················· | ···················· | |
| G | 5572 hCoV-19/Australia/VIC3272/2020\|EPI_ISL_520951\|2020 | ···················· | ···················· | ···················· | ···················· | ···················· | ···················· | |
| G | 5573 hCoV-19/Australia/VIC3273/2020\|EPI_ISL_520952\|2020 | ···················· | ···················· | ···················· | ···················· | ···················· | ···················· | |
| G | 5574 hCoV-19/Australia/VIC3275/2020\|EPI_ISL_520954\|2020 | ···················· | ···················· | ···················· | ···················· | ···················· | ···················· | |
| G | 5575 hCoV-19/Australia/VIC3276/2020\|EPI_ISL_520955\|2020 | ···················· | ···················· | ···················· | ···················· | ···················· | ···················· | |
| G | 5576 hCoV-19/Australia/VIC3277/2020\|EPI_ISL_520956\|2020 | ···················· | ···················· | ···················· | ···················· | ···················· | ···················· | |
| G | 5577 hCoV-19/Australia/VIC3278/2020\|EPI_ISL_520957\|2020 | ···················· | ···················· | ···················· | ···················· | ···················· | ···················· | |
| G | 5578 hCoV-19/Australia/VIC3279/2020\|EPI_ISL_520958\|2020 | ···················· | ···················· | ···················· | ···················· | ···················· | ···················· | |
| G | 5579 hCoV-19/Australia/VIC3280/2020\|EPI_ISL_520959\|2020 | ···················· | ···················· | ···················· | ···················· | ···················· | ···················· | |
| G | 5580 hCoV-19/Australia/VIC3281/2020\|EPI_ISL_520960\|2020 | ···················· | ···················· | ···················· | ···················· | ···················· | ···················· | |
| G | 5581 hCoV-19/Australia/VIC3282/2020\|EPI_ISL_520961\|2020 | ···················· | ···················· | ···················· | ···················· | ···················· | ···················· | |
| G | 5582 hCoV-19/Netherlands/GE-EMC-178/2020\|EPI_ISL_523160 | ···················· | ···················· | ···················· | ···················· | ···················· | ···················· | |
| G | 5583 hCoV-19/Netherlands/GE-EMC-179/2020\|EPI_ISL_523161 | ···················· | ···················· | ···················· | ···················· | ···················· | ···················· | |
| G | 5584 hCoV-19/Netherlands/GE-EMC-185/2020\|EPI_ISL_523167 | ···················· | ···················· | ···················· | ···················· | ···················· | ···················· | |
| G | 5585 hCoV-19/Netherlands/GE-EMC-186/2020\|EPI_ISL_523168 | ···················· | ···················· | ···················· | ···················· | ···················· | ···················· | |
| G | 5586 hCoV-19/Netherlands/GE-EMC-188/2020\|EPI_ISL_523170 | ···················· | ···················· | ···················· | ···················· | ···················· | ···················· | |
| G | 5587 hCoV-19/Netherlands/GE-EMC-189/2020\|EPI_ISL_523171 | ···················· | ···················· | ···················· | ···················· | ···················· | ···················· | |
| G | 5588 hCoV-19/Netherlands/GE-EMC-190/2020\|EPI_ISL_523172 | ···················· | ···················· | ···················· | ···················· | ···················· | ···················· | |

<table>
<tr><td>G</td><td>5589 hCoV-19/Netherlands/GE-EMC-191/2020|EPI_ISL_523173</td><td>····················</td><td>····················</td><td>·····················</td><td>····················</td><td>····················</td><td>····················</td><td></td><td></td></tr>
<tr><td>G</td><td>5590 hCoV-19/Netherlands/GE-EMC-192/2020|EPI_ISL_523174</td><td>····················</td><td>····················</td><td>·····················</td><td>····················</td><td>····················</td><td>····················</td><td></td><td></td></tr>
<tr><td>G</td><td>5591 hCoV-19/Netherlands/GE-EMC-193/2020|EPI_ISL_523175</td><td>····················</td><td>····················</td><td>·····················</td><td>····················</td><td>····················</td><td>····················</td><td></td><td></td></tr>
<tr><td>G</td><td>5592 hCoV-19/Netherlands/GE-EMC-194/2020|EPI_ISL_523176</td><td>····················</td><td>····················</td><td>·····················</td><td>····················</td><td>····················</td><td>····················</td><td></td><td></td></tr>
<tr><td>G</td><td>5593 hCoV-19/Netherlands/GE-EMC-195/2020|EPI_ISL_523177</td><td>····················</td><td>····················</td><td>·····················</td><td>····················</td><td>····················</td><td>····················</td><td></td><td></td></tr>
<tr><td>G</td><td>5594 hCoV-19/Netherlands/GE-EMC-196/2020|EPI_ISL_523178</td><td>····················</td><td>····················</td><td>·····················</td><td>····················</td><td>····················</td><td>····················</td><td></td><td></td></tr>
<tr><td>G</td><td>5595 hCoV-19/Netherlands/GE-EMC-197/2020|EPI_ISL_523179</td><td>····················</td><td>····················</td><td>·····················</td><td>····················</td><td>····················</td><td>····················</td><td></td><td></td></tr>
<tr><td>G</td><td>5596 hCoV-19/Netherlands/GE-EMC-198/2020|EPI_ISL_523180</td><td>····················</td><td>····················</td><td>·····················</td><td>····················</td><td>····················</td><td>····················</td><td></td><td></td></tr>
<tr><td>G</td><td>5597 hCoV-19/Netherlands/GE-EMC-199/2020|EPI_ISL_523181</td><td>····················</td><td>····················</td><td>·····················</td><td>····················</td><td>····················</td><td>····················</td><td></td><td></td></tr>
<tr><td>G</td><td>5598 hCoV-19/Netherlands/GE-EMC-200/2020|EPI_ISL_523182</td><td>····················</td><td>····················</td><td>·····················</td><td>····················</td><td>····················</td><td>····················</td><td></td><td></td></tr>
<tr><td>G</td><td>5599 hCoV-19/Netherlands/GE-EMC-201/2020|EPI_ISL_523183</td><td>····················</td><td>····················</td><td>·····················</td><td>····················</td><td>····················</td><td>····················</td><td></td><td></td></tr>
<tr><td>G</td><td>5600 hCoV-19/Netherlands/GE-EMC-202/2020|EPI_ISL_523184</td><td>····················</td><td>····················</td><td>·····················</td><td>····················</td><td>····················</td><td>····················</td><td></td><td></td></tr>
<tr><td>G</td><td>5601 hCoV-19/Netherlands/GE-EMC-203/2020|EPI_ISL_523185</td><td>····················</td><td>····················</td><td>·····················</td><td>····················</td><td>····················</td><td>····················</td><td></td><td></td></tr>
<tr><td>G</td><td>5602 hCoV-19/Netherlands/GE-EMC-204/2020|EPI_ISL_523186</td><td>····················</td><td>····················</td><td>·····················</td><td>····················</td><td>····················</td><td>····················</td><td></td><td></td></tr>
<tr><td>G</td><td>5603 hCoV-19/Netherlands/GE-EMC-205/2020|EPI_ISL_523187</td><td>····················</td><td>····················</td><td>·····················</td><td>····················</td><td>····················</td><td>····················</td><td></td><td></td></tr>
<tr><td>G</td><td>5604 hCoV-19/Netherlands/GE-EMC-206/2020|EPI_ISL_523188</td><td>····················</td><td>····················</td><td>·····················</td><td>····················</td><td>····················</td><td>····················</td><td></td><td></td></tr>
<tr><td>G</td><td>5605 hCoV-19/Netherlands/GE-EMC-207/2020|EPI_ISL_523189</td><td>····················</td><td>····················</td><td>·····················</td><td>····················</td><td>····················</td><td>····················</td><td></td><td></td></tr>
<tr><td>G</td><td>5606 hCoV-19/Netherlands/GE-EMC-209/2020|EPI_ISL_523191</td><td>····················</td><td>····················</td><td>·····················</td><td>····················</td><td>····················</td><td>····················</td><td></td><td></td></tr>
<tr><td>G</td><td>5607 hCoV-19/Netherlands/GE-EMC-210/2020|EPI_ISL_523192</td><td>····················</td><td>····················</td><td>·····················</td><td>····················</td><td>····················</td><td>····················</td><td></td><td></td></tr>
<tr><td>G</td><td>5608 hCoV-19/Netherlands/GE-EMC-211/2020|EPI_ISL_523193</td><td>····················</td><td>····················</td><td>·····················</td><td>····················</td><td>····················</td><td>····················</td><td></td><td></td></tr>
<tr><td>G</td><td>5609 hCoV-19/Netherlands/GE-EMC-213/2020|EPI_ISL_523195</td><td>····················</td><td>····················</td><td>·····················</td><td>····················</td><td>····················</td><td>····················</td><td></td><td></td></tr>
<tr><td>G</td><td>5610 hCoV-19/Netherlands/GE-EMC-214/2020|EPI_ISL_523196</td><td>····················</td><td>····················</td><td>·····················</td><td>····················</td><td>····················</td><td>····················</td><td></td><td></td></tr>
<tr><td>G</td><td>5611 hCoV-19/Netherlands/GE-EMC-215/2020|EPI_ISL_523197</td><td>····················</td><td>····················</td><td>·····················</td><td>····················</td><td>····················</td><td>····················</td><td></td><td></td></tr>
<tr><td>G</td><td>5612 hCoV-19/Netherlands/GE-EMC-217/2020|EPI_ISL_523199</td><td>····················</td><td>····················</td><td>·····················</td><td>····················</td><td>····················</td><td>····················</td><td></td><td></td></tr>
<tr><td>G</td><td>5613 hCoV-19/USA/UT-08684/2020|EPI_ISL_524405|2020-05-0</td><td>····················</td><td>····················</td><td>·····················</td><td>····················</td><td>····················</td><td>····················</td><td></td><td></td></tr>
<tr><td>G</td><td>5614 hCoV-19/USA/UT-08688/2020|EPI_ISL_524408|2020-05-1</td><td>····················</td><td>····················</td><td>·····················</td><td>····················</td><td>····················</td><td>····················</td><td></td><td></td></tr>
<tr><td>G</td><td>5615 hCoV-19/England/BRIS-133892/2020|EPI_ISL_524490|20</td><td>····················</td><td>····················</td><td>·········T···········</td><td>····················</td><td>····················</td><td>····················</td><td></td><td></td></tr>
<tr><td>G</td><td>5616 hCoV-19/England/BRIS-1338CF/2020|EPI_ISL_524491|20</td><td>····················</td><td>····················</td><td>·····················</td><td>····················</td><td>····················</td><td>····················</td><td></td><td></td></tr>
<tr><td>G</td><td>5617 hCoV-19/England/BRIS-133838/2020|EPI_ISL_524492|20</td><td>····················</td><td>····················</td><td>·········T···········</td><td>····················</td><td>····················</td><td>····················</td><td></td><td></td></tr>
<tr><td>G</td><td>5618 hCoV-19/Scotland/GCVR-170DA9/2020|EPI_ISL_524494|2</td><td>····················</td><td>····A···············</td><td>·····················</td><td>····················</td><td>····················</td><td>····················</td><td></td><td></td></tr>
<tr><td>G</td><td>5619 hCoV-19/England/CAMB-7E42F/2020|EPI_ISL_524495|202</td><td>····················</td><td>····················</td><td>·····················</td><td>····················</td><td>····················</td><td>····················</td><td></td><td></td></tr>
<tr><td>G</td><td>5620 hCoV-19/England/CAMB-80369/2020|EPI_ISL_524497|202</td><td>····················</td><td>····················</td><td>·····················</td><td>····················</td><td>····················</td><td>····················</td><td></td><td></td></tr>
<tr><td>G</td><td>5621 hCoV-19/England/CAMB-8030F/2020|EPI_ISL_524498|202</td><td>····················</td><td>····················</td><td>·····················</td><td>····················</td><td>····················</td><td>····················</td><td></td><td></td></tr>
<tr><td>G</td><td>5622 hCoV-19/USA/WA-S2747/2020|EPI_ISL_525737|2020-07-1</td><td>····················</td><td>····················</td><td>·····················</td><td>····················</td><td>····················</td><td>····················</td><td></td><td></td></tr>
<tr><td>G</td><td>5623 hCoV-19/USA/WA-S2748/2020|EPI_ISL_525738|2020-07-1</td><td>····················</td><td>·A··················</td><td>·····················</td><td>····················</td><td>····················</td><td>····················</td><td></td><td></td></tr>
<tr><td>G</td><td>5624 hCoV-19/USA/WA-S2749/2020|EPI_ISL_525739|2020-07-2</td><td>····················</td><td>····················</td><td>·····················</td><td>····················</td><td>····················</td><td>····················</td><td></td><td></td></tr>
<tr><td>G</td><td>5625 hCoV-19/Switzerland/BS-UHB-42189220/2020|EPI_ISL_5</td><td>····················</td><td>····················</td><td>·····················</td><td>····················</td><td>····················</td><td>····················</td><td></td><td></td></tr>
<tr><td>G</td><td>5626 hCoV-19/Switzerland/BS-UHB-42189239/2020|EPI_ISL_5</td><td>····················</td><td>·A··················</td><td>·····················</td><td>····················</td><td>····················</td><td>····················</td><td></td><td></td></tr>
<tr><td>G</td><td>5627 hCoV-19/Switzerland/BL-UHB-42189257/2020|EPI_ISL_5</td><td>····················</td><td>····················</td><td>·····················</td><td>····················</td><td>····················</td><td>····················</td><td></td><td></td></tr>
<tr><td>G</td><td>5628 hCoV-19/Switzerland/BS-UHB-42189386/2020|EPI_ISL_5</td><td>····················</td><td>····················</td><td>·····················</td><td>····················</td><td>····················</td><td>····················</td><td></td><td></td></tr>
<tr><td>G</td><td>5629 hCoV-19/Switzerland/BL-UHB-42189451/2020|EPI_ISL_5</td><td>····················</td><td>····················</td><td>·····················</td><td>····················</td><td>····················</td><td>····················</td><td></td><td></td></tr>
<tr><td>G</td><td>5630 hCoV-19/Switzerland/BS-UHB-42189582/2020|EPI_ISL_5</td><td>····················</td><td>····················</td><td>·····················</td><td>····················</td><td>····················</td><td>····················</td><td></td><td></td></tr>
<tr><td>G</td><td>5631 hCoV-19/Switzerland/BS-UHB-42189643/2020|EPI_ISL_5</td><td>····················</td><td>····················</td><td>·····················</td><td>····················</td><td>····················</td><td>····················</td><td></td><td></td></tr>
<tr><td>G</td><td>5632 hCoV-19/Switzerland/BL-UHB-42189647/2020|EPI_ISL_5</td><td>····················</td><td>····················</td><td>·····················</td><td>····················</td><td>····················</td><td>····················</td><td></td><td></td></tr>
<tr><td>G</td><td>5633 hCoV-19/England/LIVE-A8592/2020|EPI_ISL_517183|202</td><td>····················</td><td>····················</td><td>·····················</td><td>····················</td><td>····················</td><td>····················</td><td></td><td></td></tr>
<tr><td>G</td><td>5634 hCoV-19/England/LIVE-A8617/2020|EPI_ISL_517190|202</td><td>····················</td><td>····················</td><td>·····················</td><td>····················</td><td>····················</td><td>····················</td><td></td><td></td></tr>
<tr><td>G</td><td>5635 hCoV-19/England/LIVE-A8626/2020|EPI_ISL_517191|202</td><td>····················</td><td>····················</td><td>·····················</td><td>····················</td><td>····················</td><td>····················</td><td></td><td></td></tr>
<tr><td>G</td><td>5636 hCoV-19/England/LIVE-A8635/2020|EPI_ISL_517192|202</td><td>····················</td><td>····················</td><td>·····················</td><td>····················</td><td>····················</td><td>····················</td><td></td><td></td></tr>
<tr><td>G</td><td>5637 hCoV-19/England/LIVE-A8644/2020|EPI_ISL_517193|202</td><td>····················</td><td>····················</td><td>·····················</td><td>····················</td><td>····················</td><td>····················</td><td></td><td></td></tr>
<tr><td>G</td><td>5638 hCoV-19/England/LIVE-A8662/2020|EPI_ISL_517195|202</td><td>····················</td><td>····················</td><td>·····················</td><td>····················</td><td>····················</td><td>····················</td><td></td><td></td></tr>
<tr><td>G</td><td>5642 hCoV-19/England/LIVE-A9ADE/2020|EPI_ISL_517320|202</td><td>····················</td><td>····················</td><td>·····················</td><td>····················</td><td>····················</td><td>····················</td><td></td><td></td></tr>
<tr><td>G</td><td>5643 hCoV-19/England/LIVE-A9AFC/2020|EPI_ISL_517322|202</td><td>····················</td><td>····················</td><td>·····················</td><td>····················</td><td>·······NN···········</td><td>····················</td><td></td><td></td></tr>
<tr><td>G</td><td>5644 hCoV-19/England/LIVE-A9B08/2020|EPI_ISL_517323|202</td><td>····················</td><td>····················</td><td>·····················</td><td>····················</td><td>····················</td><td>····················</td><td></td><td></td></tr>
<tr><td>G</td><td>5645 hCoV-19/England/LIVE-A9B17/2020|EPI_ISL_517324|202</td><td>····················</td><td>····················</td><td>·····················</td><td>····················</td><td>····················</td><td>····················</td><td></td><td></td></tr>
<tr><td>G</td><td>5647 hCoV-19/England/LIVE-A9B35/2020|EPI_ISL_517326|202</td><td>····················</td><td>····················</td><td>·····················</td><td>····················</td><td>····················</td><td>····················</td><td></td><td></td></tr>
<tr><td>G</td><td>5648 hCoV-19/England/LIVE-A9B44/2020|EPI_ISL_517327|202</td><td>····················</td><td>····················</td><td>·····················</td><td>····················</td><td>····················</td><td>····················</td><td></td><td></td></tr>
<tr><td>G</td><td>5649 hCoV-19/England/LIVE-A9B53/2020|EPI_ISL_517328|202</td><td>····················</td><td>····················</td><td>·····················</td><td>····················</td><td>····················</td><td>····················</td><td></td><td></td></tr>
<tr><td>G</td><td>5650 hCoV-19/England/LIVE-A9B71/2020|EPI_ISL_517329|202</td><td>····················</td><td>···················A</td><td>·····················</td><td>····················</td><td>····················</td><td>····················</td><td>1</td><td></td></tr>
<tr><td>G</td><td>5651 hCoV-19/England/LIVE-A9B80/2020|EPI_ISL_517330|202</td><td>····················</td><td>····················</td><td>·····················</td><td>····················</td><td>····················</td><td>····················</td><td></td><td></td></tr>
<tr><td>G</td><td>5652 hCoV-19/England/LIVE-A9B9F/2020|EPI_ISL_517331|202</td><td>····················</td><td>····················</td><td>·····················</td><td>····················</td><td>····················</td><td>····················</td><td></td><td></td></tr>
<tr><td>G</td><td>5654 hCoV-19/England/LIVE-A9BBD/2020|EPI_ISL_517333|202</td><td>····················</td><td>····················</td><td>·····················</td><td>····················</td><td>····················</td><td>····················</td><td></td><td></td></tr>
<tr><td>G</td><td>5656 hCoV-19/England/LIVE-A9BEA/2020|EPI_ISL_517335|202</td><td>····················</td><td>····················</td><td>·····················</td><td>····················</td><td>····················</td><td>····················</td><td></td><td></td></tr>
<tr><td>G</td><td>5657 hCoV-19/England/LIVE-A9BF9/2020|EPI_ISL_517336|202</td><td>····················</td><td>····················</td><td>·····················</td><td>····················</td><td>····················</td><td>····················</td><td></td><td></td></tr>
<tr><td>G</td><td>5658 hCoV-19/England/LIVE-A9C05/2020|EPI_ISL_517337|202</td><td>····················</td><td>····················</td><td>·····················</td><td>····················</td><td>····················</td><td>····················</td><td></td><td></td></tr>
<tr><td>G</td><td>5659 hCoV-19/England/LIVE-A9C14/2020|EPI_ISL_517338|202</td><td>····················</td><td>····················</td><td>·····················</td><td>····················</td><td>····················</td><td>····················</td><td></td><td></td></tr>
<tr><td>G</td><td>5660 hCoV-19/England/LIVE-A9C23/2020|EPI_ISL_517339|202</td><td>····················</td><td>····················</td><td>·····················</td><td>····················</td><td>····················</td><td>····················</td><td></td><td></td></tr>
<tr><td>G</td><td>5662 hCoV-19/England/LIVE-A9C41/2020|EPI_ISL_517341|202</td><td>····················</td><td>····················</td><td>·····················</td><td>····················</td><td>····················</td><td>····················</td><td></td><td></td></tr>
<tr><td>G</td><td>5664 hCoV-19/England/LIVE-A9C7E/2020|EPI_ISL_517344|202</td><td>····················</td><td>····················</td><td>·····················</td><td>····················</td><td>·······NNN··········</td><td>····················</td><td></td><td>1</td></tr>
<tr><td>G</td><td>5665 hCoV-19/England/LIVE-A9C8D/2020|EPI_ISL_517345|202</td><td>····················</td><td>····················</td><td>·····················</td><td>····················</td><td>····················</td><td>····················</td><td></td><td></td></tr>
<tr><td>G</td><td>5666 hCoV-19/England/LIVE-A9C9C/2020|EPI_ISL_517346|202</td><td>····················</td><td>····················</td><td>·····················</td><td>····················</td><td>·······NN···········</td><td>····················</td><td></td><td></td></tr>
<tr><td>G</td><td>5667 hCoV-19/England/LIVE-A9CAB/2020|EPI_ISL_517347|202</td><td>····················</td><td>····················</td><td>·····················</td><td>····················</td><td>····················</td><td>····················</td><td></td><td></td></tr>
<tr><td>G</td><td>5668 hCoV-19/England/LIVE-A9CBA/2020|EPI_ISL_517348|202</td><td>····················</td><td>····················</td><td>·····················</td><td>····················</td><td>····················</td><td>····················</td><td></td><td></td></tr>
<tr><td>G</td><td>5669 hCoV-19/England/LIVE-A9CC9/2020|EPI_ISL_517349|202</td><td>····················</td><td>····················</td><td>·····················</td><td>····················</td><td>····················</td><td>····················</td><td></td><td></td></tr>
<tr><td>G</td><td>5670 hCoV-19/England/LIVE-A9CD8/2020|EPI_ISL_517350|202</td><td>····················</td><td>····················</td><td>·····················</td><td>····················</td><td>····················</td><td>····················</td><td></td><td></td></tr>
<tr><td>G</td><td>5671 hCoV-19/England/LIVE-A9CE7/2020|EPI_ISL_517351|202</td><td>····················</td><td>····················</td><td>·····················</td><td>····················</td><td>····················</td><td>····················</td><td></td><td></td></tr>
<tr><td>G</td><td>5672 hCoV-19/England/LIVE-A9CF6/2020|EPI_ISL_517352|202</td><td>····················</td><td>····················</td><td>·····················</td><td>····················</td><td>····················</td><td>····················</td><td></td><td></td></tr>
<tr><td>G</td><td>5673 hCoV-19/England/LIVE-A9D02/2020|EPI_ISL_517353|202</td><td>····················</td><td>····················</td><td>·····················</td><td>····················</td><td>····················</td><td>····················</td><td></td><td></td></tr>
<tr><td>G</td><td>5674 hCoV-19/England/LIVE-A9D11/2020|EPI_ISL_517354|202</td><td>····················</td><td>····················</td><td>·····················</td><td>····················</td><td>·······NN···········</td><td>····················</td><td></td><td></td></tr>
<tr><td>G</td><td>5676 hCoV-19/England/LIVE-A9D3F/2020|EPI_ISL_517356|202</td><td>····················</td><td>····················</td><td>·····················</td><td>····················</td><td>····················</td><td>····················</td><td></td><td></td></tr>
<tr><td>G</td><td>5677 hCoV-19/England/LIVE-A9D4E/2020|EPI_ISL_517357|202</td><td>····················</td><td>····················</td><td>·····················</td><td>····················</td><td>····················</td><td>····················</td><td></td><td></td></tr>
</table>

| | | | | | | | | |
|---|---|---|---|---|---|---|---|---|
| G | 5678 hCoV-19/England/LIVE-A9D5D/2020\|EPI_ISL_517358\|202 | .................... | .................... | .................... | .................... | ......NN............ | .................... | |
| G | 5679 hCoV-19/England/LIVE-A9D8A/2020\|EPI_ISL_517359\|202 | .................... | .................... | .................... | .................... | .................... | .................... | |
| G | 5680 hCoV-19/England/LIVE-A9D99/2020\|EPI_ISL_517360\|202 | .................... | ...................A | .................... | .................... | .................... | .................... | 1 |
| G | 5681 hCoV-19/England/LIVE-A9DA8/2020\|EPI_ISL_517361\|202 | .................... | .................... | .................... | .................... | .................... | .................... | |
| G | 5682 hCoV-19/England/LIVE-A9DB7/2020\|EPI_ISL_517362\|202 | .................... | .................... | .................... | .................... | .................... | .................... | |
| G | 5683 hCoV-19/England/LIVE-A9DC6/2020\|EPI_ISL_517363\|202 | .................... | .................... | .................... | .................... | .................... | .................... | |
| G | 5684 hCoV-19/England/LIVE-A9DD5/2020\|EPI_ISL_517364\|202 | .................... | .................... | .................... | .................... | .................... | .................... | |
| G | 5685 hCoV-19/England/LIVE-A9DE4/2020\|EPI_ISL_517365\|202 | .................... | .................... | .................... | .................... | .................... | .................... | |
| G | 5686 hCoV-19/England/LIVE-A9DF3/2020\|EPI_ISL_517366\|202 | .................... | .................... | .................... | .................... | .................... | .................... | |
| G | 5687 hCoV-19/England/LIVE-A9E0F/2020\|EPI_ISL_517367\|202 | .................... | .................... | .................... | .................... | .................... | .................... | |
| G | 5688 hCoV-19/England/LIVE-A9E1E/2020\|EPI_ISL_517368\|202 | .................... | .................... | .................... | .................... | .................... | .................... | |
| G | 5689 hCoV-19/England/LIVE-A9E4B/2020\|EPI_ISL_517369\|202 | .................... | .................... | .................... | .................... | ......NN............ | .................... | |
| G | 5692 hCoV-19/England/LIVE-A9F1B/2020\|EPI_ISL_517373\|202 | .................... | .................... | .................... | .................... | ......NN............ | .................... | |
| G | 5693 hCoV-19/England/LIVE-A9F84/2020\|EPI_ISL_517380\|202 | .................... | .................... | .................... | .................... | .................... | .................... | |
| G | 5694 hCoV-19/England/LIVE-A9FDF/2020\|EPI_ISL_517385\|202 | .................... | .................... | .................... | .................... | .................... | .................... | |
| G | 5695 hCoV-19/England/LIVE-AA080/2020\|EPI_ISL_517396\|202 | .................... | .................... | .................... | .................... | .................... | .................... | |
| G | 5696 hCoV-19/England/LIVE-AA09F/2020\|EPI_ISL_517397\|202 | .................... | .................... | .................... | .................... | .................... | .................... | |
| G | 5697 hCoV-19/England/LIVE-AA0BD/2020\|EPI_ISL_517399\|202 | .................... | .................... | .................... | .................... | .................... | .................... | |
| G | 5698 hCoV-19/Australia/VIC5858/2020\|EPI_ISL_518651\|2020 | .................... | .................... | .................... | .................... | .................... | .................... | |
| G | 5699 hCoV-19/Australia/VIC5859/2020\|EPI_ISL_518652\|2020 | .................... | .................... | .................... | .................... | .................... | .................... | |
| G | 5700 hCoV-19/Australia/VIC5860/2020\|EPI_ISL_518653\|2020 | .................... | .................... | .................... | .................... | .................... | .................... | |
| G | 5701 hCoV-19/Australia/VIC5861/2020\|EPI_ISL_518654\|2020 | .................... | .................... | .................... | .................... | .................... | .................... | |
| G | 5702 hCoV-19/Australia/VIC5862/2020\|EPI_ISL_518655\|2020 | .................... | .................... | .................... | .................... | .................... | .................... | |
| G | 5703 hCoV-19/Australia/VIC5863/2020\|EPI_ISL_518656\|2020 | .................... | .................... | .................... | .................... | .................... | .................... | |
| G | 5704 hCoV-19/Australia/VIC5864/2020\|EPI_ISL_518657\|2020 | .................... | .................... | .................... | .................... | .................... | .................... | |
| G | 5705 hCoV-19/Australia/VIC5867/2020\|EPI_ISL_518658\|2020 | .................... | .................... | .................... | .................... | .................... | .................... | |
| G | 5706 hCoV-19/Australia/VIC5869/2020\|EPI_ISL_518659\|2020 | .................... | .................... | .................... | .................... | .................... | .................... | |
| G | 5707 hCoV-19/Australia/VIC5870/2020\|EPI_ISL_518660\|2020 | .................... | .................... | .................... | .................... | .................... | .................... | |
| G | 5708 hCoV-19/Australia/VIC5876/2020\|EPI_ISL_518662\|2020 | .................... | .................... | .................... | .................... | .................... | .................... | |
| G | 5709 hCoV-19/Australia/VIC5881/2020\|EPI_ISL_518664\|2020 | .................... | .................... | .................... | .................... | .................... | .................... | |
| G | 5710 hCoV-19/Australia/VIC5882/2020\|EPI_ISL_518665\|2020 | .................... | .................... | .................... | .................... | .................... | .................... | |
| G | 5711 hCoV-19/Australia/VIC5883/2020\|EPI_ISL_518666\|2020 | .................... | .................... | .................... | .................... | .................... | .................... | |
| G | 5712 hCoV-19/Australia/VIC5884/2020\|EPI_ISL_518667\|2020 | .................... | .................... | .................... | .................... | .................... | .................... | |
| G | 5713 hCoV-19/Australia/VIC5886/2020\|EPI_ISL_518669\|2020 | .................... | .................... | .................... | .................... | .................... | .................... | |
| G | 5714 hCoV-19/Australia/VIC5887/2020\|EPI_ISL_518670\|2020 | .................... | .................... | .................... | .................... | .................... | .................... | |
| G | 5715 hCoV-19/Australia/VIC5889/2020\|EPI_ISL_518671\|2020 | .................... | .................... | .................... | .................... | .................... | .................... | |
| G | 5716 hCoV-19/Australia/VIC5890/2020\|EPI_ISL_518672\|2020 | .................... | .................... | .................... | .................... | .................... | .................... | |
| G | 5717 hCoV-19/Australia/VIC5891/2020\|EPI_ISL_518673\|2020 | .................... | .................... | .................... | .................... | .................... | .................... | |
| G | 5718 hCoV-19/Australia/VIC5892/2020\|EPI_ISL_518674\|2020 | .................... | .................... | .................... | .................... | .................... | .................... | |
| G | 5719 hCoV-19/Australia/VIC5893/2020\|EPI_ISL_518675\|2020 | .................... | .................... | .................... | .................... | .................... | .................... | |
| G | 5720 hCoV-19/Australia/VIC5894/2020\|EPI_ISL_518676\|2020 | .................... | .................... | .................... | .................... | .................... | .................... | |
| G | 5721 hCoV-19/Australia/VIC5896/2020\|EPI_ISL_518677\|2020 | .................... | .................... | .................... | .................... | .................... | .................... | |
| G | 5722 hCoV-19/Australia/VIC5897/2020\|EPI_ISL_518678\|2020 | .................... | .................... | .................... | .................... | .................... | .................... | |
| G | 5723 hCoV-19/Australia/VIC5898/2020\|EPI_ISL_518679\|2020 | .................... | .................... | .................... | .................... | .................... | .................... | |
| G | 5724 hCoV-19/Australia/VIC5899/2020\|EPI_ISL_518680\|2020 | .................... | .................... | .................... | .................... | .................... | .................... | |
| G | 5725 hCoV-19/Australia/VIC5900/2020\|EPI_ISL_518681\|2020 | .................... | .................... | .................... | .................... | .................... | .................... | |
| G | 5726 hCoV-19/Australia/VIC5902/2020\|EPI_ISL_518682\|2020 | .................... | .................... | .................... | .................... | .................... | .................... | |
| G | 5727 hCoV-19/Australia/VIC5903/2020\|EPI_ISL_518683\|2020 | .................... | .................... | .................... | .................... | .................... | .................... | |
| G | 5728 hCoV-19/Australia/VIC5904/2020\|EPI_ISL_518684\|2020 | .................... | .................... | .................... | .................... | .................... | .................... | |
| G | 5729 hCoV-19/Australia/VIC5906/2020\|EPI_ISL_518685\|2020 | .................... | .................... | .................... | .................... | .................... | .................... | |
| G | 5730 hCoV-19/Australia/VIC5907/2020\|EPI_ISL_518686\|2020 | .................... | .................... | .................... | .................... | .................... | .................... | |
| G | 5731 hCoV-19/Australia/VIC5908/2020\|EPI_ISL_518687\|2020 | .................... | .................... | .................... | .................... | .................... | .................... | |
| G | 5732 hCoV-19/Australia/VIC5909/2020\|EPI_ISL_518688\|2020 | .................... | .................... | .................... | .................... | .................... | .................... | |
| G | 5733 hCoV-19/Australia/VIC5910/2020\|EPI_ISL_518689\|2020 | .................... | .................... | .................... | .................... | .................... | .................... | |
| G | 5734 hCoV-19/Australia/VIC5911/2020\|EPI_ISL_518690\|2020 | .................... | .................... | .................... | .................... | .................... | .................... | |
| G | 5735 hCoV-19/Australia/VIC5912/2020\|EPI_ISL_518691\|2020 | .................... | .................... | .................... | .................... | .................... | .................... | |
| G | 5736 hCoV-19/Australia/VIC5913/2020\|EPI_ISL_518692\|2020 | .................... | .................... | .................... | .................... | .................... | .................... | |
| G | 5738 hCoV-19/Australia/VIC5915/2020\|EPI_ISL_518694\|2020 | .................... | .................... | .................... | .................... | .................... | .................... | |
| G | 5739 hCoV-19/Australia/VIC5916/2020\|EPI_ISL_518695\|2020 | .................... | .................... | .................... | .................... | .................... | .................... | |
| G | 5740 hCoV-19/Australia/VIC5917/2020\|EPI_ISL_518696\|2020 | .................... | .................... | .................... | .................... | .................... | .................... | |
| G | 5741 hCoV-19/Australia/VIC5918/2020\|EPI_ISL_518697\|2020 | .................... | .................... | .................... | .................... | .................... | .................... | |
| G | 5742 hCoV-19/Australia/VIC5919/2020\|EPI_ISL_518698\|2020 | .................... | .................... | .................... | .................... | .................... | .................... | |
| G | 5743 hCoV-19/Australia/VIC5920/2020\|EPI_ISL_518699\|2020 | .................... | .................... | .................... | .................... | .................... | .................... | |
| G | 5744 hCoV-19/Australia/VIC5215/2020\|EPI_ISL_519980\|2020 | .................... | .................... | .................... | .................... | .................... | .................... | |
| G | 5745 hCoV-19/Australia/VIC5216/2020\|EPI_ISL_519981\|2020 | .................... | .................... | .................... | .................... | .................... | .................... | |
| G | 5746 hCoV-19/Australia/VIC5217/2020\|EPI_ISL_519982\|2020 | .................... | .................... | .................... | .................... | .................... | .................... | |
| G | 5747 hCoV-19/Australia/VIC5218/2020\|EPI_ISL_519983\|2020 | .................... | .................... | .................... | .................... | .................... | .................... | |
| G | 5748 hCoV-19/Australia/VIC5219/2020\|EPI_ISL_519984\|2020 | .................... | .................... | .................... | .................... | .................... | .................... | |
| G | 5749 hCoV-19/Australia/VIC5220/2020\|EPI_ISL_519985\|2020 | .................... | .................... | .................... | .................... | .................... | .................... | |
| G | 5750 hCoV-19/Australia/VIC5221/2020\|EPI_ISL_519986\|2020 | .................... | .................... | .................... | .................... | .................... | .................... | |
| G | 5751 hCoV-19/Australia/VIC5222/2020\|EPI_ISL_519987\|2020 | .................... | .................... | .................... | .................... | .................... | .................... | |
| G | 5752 hCoV-19/Australia/VIC5223/2020\|EPI_ISL_519988\|2020 | .................... | .................... | .................... | .................... | .................... | .................... | |
| G | 5753 hCoV-19/Australia/VIC5224/2020\|EPI_ISL_519989\|2020 | .................... | .................... | .................... | .................... | .................... | .................... | |
| G | 5754 hCoV-19/Australia/VIC5225/2020\|EPI_ISL_519990\|2020 | .................... | .................... | .................... | .................... | .................... | .................... | |
| G | 5755 hCoV-19/Australia/VIC5226/2020\|EPI_ISL_519991\|2020 | .................... | .................... | .................... | .................... | .................... | .................... | |
| G | 5756 hCoV-19/Australia/VIC5227/2020\|EPI_ISL_519992\|2020 | .................... | .................... | .................... | .................... | .................... | .................... | |
| G | 5757 hCoV-19/Australia/VIC5228/2020\|EPI_ISL_519993\|2020 | .................... | .................... | .................... | .................... | .................... | .................... | |
| G | 5758 hCoV-19/Australia/VIC5229/2020\|EPI_ISL_519994\|2020 | .................... | .................... | .................... | .................... | .................... | .................... | |
| G | 5759 hCoV-19/Australia/VIC5230/2020\|EPI_ISL_519995\|2020 | .................... | .................... | .................... | .................... | .................... | .................... | |
| G | 5760 hCoV-19/Australia/VIC5231/2020\|EPI_ISL_519996\|2020 | .................... | .................... | .................... | .................... | .................... | .................... | |

| | | | | | | | | |
|---|---|---|---|---|---|---|---|---|
| G | 5761 hCoV-19/Australia/VIC5232/2020\|EPI_ISL_519997\|2020 | ···················· | ···················· | ···················· | ···················· | ···················· | ···················· | |
| G | 5762 hCoV-19/Australia/VIC5233/2020\|EPI_ISL_519998\|2020 | ···················· | ···················· | ···················· | ···················· | ···················· | ···················· | |
| G | 5763 hCoV-19/Australia/VIC5234/2020\|EPI_ISL_519999\|2020 | ···················· | ···················· | ···················· | ···················· | ···················· | ···················· | |
| G | 5764 hCoV-19/Australia/VIC3283/2020\|EPI_ISL_520962\|2020 | ···················· | ···················· | ···················· | ···················· | ···················· | ···················· | |
| G | 5765 hCoV-19/Australia/VIC3285/2020\|EPI_ISL_520964\|2020 | ···················· | ···················· | ···················· | ···················· | ···················· | ···················· | |
| G | 5766 hCoV-19/Australia/VIC3286/2020\|EPI_ISL_520965\|2020 | ···················· | ···················· | ···················· | ···················· | ···················· | ···················· | |
| G | 5767 hCoV-19/Australia/VIC3287/2020\|EPI_ISL_520966\|2020 | ···················· | ···················· | ···················· | ···················· | ···················· | ···················· | |
| G | 5768 hCoV-19/Australia/VIC3288/2020\|EPI_ISL_520967\|2020 | ···················· | ···················· | ···················· | ···················· | ···················· | ···················· | |
| G | 5769 hCoV-19/Australia/VIC3289/2020\|EPI_ISL_520968\|2020 | ···················· | ···················· | ···················· | ···················· | ···················· | ···················· | |
| G | 5770 hCoV-19/Australia/VIC3290/2020\|EPI_ISL_520969\|2020 | ···················· | ···················· | ···················· | ···················· | ···················· | ···················· | |
| G | 5771 hCoV-19/Australia/VIC3291/2020\|EPI_ISL_520970\|2020 | ···················· | ···················· | ···················· | ···················· | ···················· | ···················· | |
| G | 5772 hCoV-19/Australia/VIC3292/2020\|EPI_ISL_520971\|2020 | ···················· | ···················· | ···················· | ···················· | ···················· | ···················· | |
| G | 5773 hCoV-19/Australia/VIC3293/2020\|EPI_ISL_520972\|2020 | ···················· | ···················· | ···················· | ···················· | ···················· | ···················· | |
| G | 5774 hCoV-19/Australia/VIC3294/2020\|EPI_ISL_520973\|2020 | ···················· | ···················· | ···················· | ···················· | ···················· | ···················· | |
| G | 5775 hCoV-19/Australia/VIC3295/2020\|EPI_ISL_520974\|2020 | ···················· | ···················· | ···················· | ···················· | ···················· | ···················· | |
| G | 5776 hCoV-19/Australia/VIC3296/2020\|EPI_ISL_520975\|2020 | ···················· | ···················· | ···················· | ···················· | ···················· | ···················· | |
| G | 5777 hCoV-19/Australia/VIC3297/2020\|EPI_ISL_520976\|2020 | ···················· | ···················· | ···················· | ···················· | ···················· | ···················· | |
| G | 5778 hCoV-19/Australia/VIC3298/2020\|EPI_ISL_520977\|2020 | ···················· | ···················· | ···················· | ···················· | ···················· | ···················· | |
| G | 5779 hCoV-19/Australia/VIC3299/2020\|EPI_ISL_520978\|2020 | ···················· | ···················· | ···················· | ···················· | ···················· | ···················· | |
| G | 5780 hCoV-19/Australia/VIC3300/2020\|EPI_ISL_520979\|2020 | ···················· | ···················· | ···················· | ···················· | ···················· | ···················· | |
| G | 5781 hCoV-19/Australia/VIC3301/2020\|EPI_ISL_520980\|2020 | ···················· | ···················· | ···················· | ···················· | ···················· | ···················· | |
| G | 5782 hCoV-19/Australia/VIC3302/2020\|EPI_ISL_520981\|2020 | ···················· | ···················· | ···················· | ···················· | ···················· | ···················· | |
| G | 5783 hCoV-19/Australia/VIC3303/2020\|EPI_ISL_520982\|2020 | ···················· | ···················· | ···················· | ···················· | ···················· | ···················· | |
| G | 5784 hCoV-19/Australia/VIC3304/2020\|EPI_ISL_520983\|2020 | ···················· | ···················· | ···················· | ···················· | ···················· | ···················· | |
| G | 5785 hCoV-19/Australia/VIC3305/2020\|EPI_ISL_520984\|2020 | ···················· | ···················· | ···················· | ···················· | ···················· | ···················· | |
| G | 5786 hCoV-19/Australia/VIC3306/2020\|EPI_ISL_520985\|2020 | ···················· | ···················· | ···················· | ···················· | ···················· | ···················· | |
| G | 5787 hCoV-19/Australia/VIC3307/2020\|EPI_ISL_520986\|2020 | ···················· | ···················· | ···················· | ···················· | ···················· | ···················· | |
| G | 5788 hCoV-19/Australia/VIC3308/2020\|EPI_ISL_520987\|2020 | ···················· | ···················· | ···················· | ···················· | NNN················· | ···················· | 1 |
| G | 5789 hCoV-19/Australia/VIC3309/2020\|EPI_ISL_520988\|2020 | ···················· | ···················· | ···················· | ···················· | ···················· | ···················· | |
| G | 5790 hCoV-19/Australia/VIC3310/2020\|EPI_ISL_520989\|2020 | ···················· | ···················· | ···················· | ···················· | ···················· | ···················· | |
| G | 5791 hCoV-19/Australia/VIC3311/2020\|EPI_ISL_520990\|2020 | ···················· | ···················· | ···················· | ···················· | ···················· | ···················· | |
| G | 5792 hCoV-19/Australia/VIC3312/2020\|EPI_ISL_520991\|2020 | ···················· | ···················· | ···················· | ···················· | ···················· | ···················· | |
| G | 5793 hCoV-19/Australia/VIC3313/2020\|EPI_ISL_520992\|2020 | ···················· | ···················· | ···················· | ···················· | ···················· | ···················· | |
| G | 5794 hCoV-19/Australia/VIC3314/2020\|EPI_ISL_520993\|2020 | ···················· | ···················· | ···················· | ···················· | ···················· | ···················· | |
| G | 5795 hCoV-19/Australia/VIC3315/2020\|EPI_ISL_520994\|2020 | ···················· | ···················· | ···················· | ···················· | ···················· | ···················· | |
| G | 5796 hCoV-19/Australia/VIC3316/2020\|EPI_ISL_520995\|2020 | ···················· | ···················· | ···················· | ···················· | ···················· | ···················· | |
| G | 5797 hCoV-19/Australia/VIC3317/2020\|EPI_ISL_520996\|2020 | ···················· | ···················· | ···················· | ···················· | ···················· | ···················· | |
| G | 5798 hCoV-19/Australia/VIC3318/2020\|EPI_ISL_520997\|2020 | ···················· | ···················· | ···················· | ···················· | ···················· | ···················· | |
| G | 5799 hCoV-19/Australia/VIC3319/2020\|EPI_ISL_520998\|2020 | ···················· | ···················· | ···················· | ···················· | ···················· | ···················· | |
| G | 5800 hCoV-19/Australia/VIC3320/2020\|EPI_ISL_520999\|2020 | ···················· | ···················· | ···················· | ···················· | ···················· | ···················· | |
| G | 5801 hCoV-19/USA/NY-Wadsworth-13260-01/2020\|EPI_ISL_525 | ···················· | ···················· | ···················· | ···················· | ···················· | ···················· | |
| G | 5802 hCoV-19/USA/NY-Wadsworth-13391-01/2020\|EPI_ISL_525 | ···················· | ···················· | ···················· | ···················· | ···················· | ···················· | |
| G | 5803 hCoV-19/USA/NY-Wadsworth-13459-01/2020\|EPI_ISL_525 | ···················· | ···················· | ···················· | ···················· | ···················· | ···················· | |
| G | 5804 hCoV-19/USA/NY-Wadsworth-13904-01/2020\|EPI_ISL_525 | ···················· | ···················· | ···················· | ···················· | ···················· | ···················· | |
| G | 5805 hCoV-19/USA/NY-Wadsworth-13972-01/2020\|EPI_ISL_525 | ···················· | ···················· | ···················· | ···················· | ···················· | ···················· | |
| G | 5806 hCoV-19/USA/NY-Wadsworth-13977-01/2020\|EPI_ISL_525 | ···················· | ···················· | ···················· | ···················· | ···················· | ···················· | |
| G | 5807 hCoV-19/USA/NY-Wadsworth-13978-01/2020\|EPI_ISL_525 | ···················· | ···················· | ···················· | ···················· | ···················· | ···················· | |
| G | 5808 hCoV-19/USA/NY-Wadsworth-14203-01/2020\|EPI_ISL_525 | ···················· | ···················· | ···················· | ···················· | ···················· | ···················· | |
| G | 5809 hCoV-19/USA/NY-Wadsworth-14368-01/2020\|EPI_ISL_525 | ···················· | ···················· | ···················· | ···················· | ···················· | ···················· | |
| G | 5810 hCoV-19/USA/NY-Wadsworth-14419-01/2020\|EPI_ISL_525 | ···················· | ···················· | ···················· | ···················· | ···················· | ···················· | |
| G | 5811 hCoV-19/USA/NY-Wadsworth-14422-01/2020\|EPI_ISL_525 | ···················· | ···················· | ···················· | ···················· | ···················· | ···················· | |
| G | 5812 hCoV-19/USA/NY-Wadsworth-14811-01/2020\|EPI_ISL_525 | ···················· | ···················· | ···················· | ···················· | ···················· | ···················· | |
| G | 5813 hCoV-19/USA/NY-Wadsworth-16307-01/2020\|EPI_ISL_525 | ···················· | ···················· | ···················· | ···················· | ···················· | ···················· | |
| G | 5814 hCoV-19/USA/VA-DCLS-0998/2020\|EPI_ISL_526900\|2020- | ···················· | ···················· | ···················· | ···················· | ···················· | ···················· | |
| G | 5815 hCoV-19/USA/VA-DCLS-1002/2020\|EPI_ISL_526901\|2020- | ···················· | ···················· | ···················· | ···················· | ···················· | ···················· | |
| G | 5816 hCoV-19/USA/VA-DCLS-1006/2020\|EPI_ISL_526902\|2020- | ···················· | ···················· | ···················· | ···················· | ···················· | ···················· | |
| G | 5817 hCoV-19/USA/VA-DCLS-1009/2020\|EPI_ISL_526903\|2020- | ········T··········· | ···················· | ···················· | ···················· | ···················· | ···················· | |
| G | 5818 hCoV-19/USA/VA-DCLS-1011/2020\|EPI_ISL_526904\|2020- | ···················· | ···················· | ···················· | ···················· | ···················· | ···················· | |
| G | 5819 hCoV-19/USA/VA-DCLS-1014/2020\|EPI_ISL_526905\|2020- | ···················· | ···················· | ···················· | ···················· | ···················· | ···················· | |
| G | 5820 hCoV-19/USA/VA-DCLS-1016/2020\|EPI_ISL_526906\|2020- | ···················· | ···················· | ···················· | ···················· | ···················· | ···················· | |
| G | 5821 hCoV-19/USA/VA-DCLS-1017/2020\|EPI_ISL_526907\|2020- | ···················· | ···················· | ···················· | ···················· | ···················· | ···················· | |
| G | 5822 hCoV-19/USA/VA-DCLS-1018/2020\|EPI_ISL_526908\|2020- | ···················· | ···················· | ···················· | ···················· | ···················· | ···················· | |
| G | 5823 hCoV-19/USA/VA-DCLS-1019/2020\|EPI_ISL_526909\|2020- | ···················· | ···················· | ···················· | ···················· | ···················· | ···················· | |
| G | 5824 hCoV-19/USA/VA-DCLS-1020/2020\|EPI_ISL_526910\|2020- | ···················· | ···················· | ···················· | ···················· | ···················· | ···················· | |
| G | 5825 hCoV-19/USA/VA-DCLS-1021/2020\|EPI_ISL_526911\|2020- | ···················· | ···················· | ···················· | ···················· | ···················· | ···················· | |
| G | 5826 hCoV-19/USA/VA-DCLS-1022/2020\|EPI_ISL_526912\|2020- | ···················· | ···················· | ···················· | ···················· | ···················· | ···················· | |
| G | 5827 hCoV-19/USA/VA-DCLS-1023/2020\|EPI_ISL_526913\|2020- | ···················· | ···················· | ···················· | ···················· | ···················· | ···················· | |
| G | 5828 hCoV-19/USA/VA-DCLS-1025/2020\|EPI_ISL_526914\|2020- | ···················· | ···················· | ···················· | ···················· | ···················· | ···················· | |
| G | 5829 hCoV-19/USA/VA-DCLS-1027/2020\|EPI_ISL_526915\|2020- | ···················· | ···················· | ···················· | ···················· | ···················· | ···················· | |
| G | 5830 hCoV-19/USA/VA-DCLS-1028/2020\|EPI_ISL_526916\|2020- | ···················· | ···················· | ···················· | ···················· | ···················· | ···················· | |
| G | 5831 hCoV-19/USA/VA-DCLS-1032/2020\|EPI_ISL_526917\|2020- | ···················· | ···················· | ···················· | ···················· | ···················· | ···················· | |
| G | 5832 hCoV-19/USA/VA-DCLS-1033/2020\|EPI_ISL_526918\|2020- | ···················· | ····A··············· | ···················· | ···················· | ···················· | ···················· | |
| G | 5833 hCoV-19/USA/VA-DCLS-1034/2020\|EPI_ISL_526919\|2020- | ···················· | ···················· | ···················· | ···················· | ···················· | ···················· | |
| G | 5834 hCoV-19/USA/VA-DCLS-1035/2020\|EPI_ISL_526920\|2020- | ···················· | ···················· | ···················· | ···················· | ···················· | ···················· | |
| G | 5835 hCoV-19/USA/VA-DCLS-1037/2020\|EPI_ISL_526921\|2020- | ···················· | ···················· | ···················· | ···················· | ···················· | ···················· | |
| G | 5836 hCoV-19/USA/VA-DCLS-1038/2020\|EPI_ISL_526922\|2020- | ···················· | ···················· | ···················· | ···················· | ···················· | ···················· | |
| G | 5837 hCoV-19/USA/VA-DCLS-1039/2020\|EPI_ISL_526923\|2020- | ···················· | ···················· | ···················· | ···················· | ···················· | ···················· | |
| G | 5838 hCoV-19/USA/VA-DCLS-1040/2020\|EPI_ISL_526924\|2020- | ···················· | ···················· | ···················· | ···················· | ···················· | ···················· | |
| G | 5839 hCoV-19/USA/VA-DCLS-1041/2020\|EPI_ISL_526925\|2020- | ···················· | ···················· | ···················· | ···················· | ···················· | ···················· | |
| G | 5840 hCoV-19/USA/VA-DCLS-1043/2020\|EPI_ISL_526926\|2020- | ···················· | ···················· | ···················· | ···················· | ···················· | ···················· | |

| | | | | | | | | | |
|---|---|---|---|---|---|---|---|---|---|
| G | 5841 hCoV-19/USA/VA-DCLS-1044/2020\|EPI_ISL_526927\|2020- | ···················· | ···················· | ···················· | ···················· | ···················· | ···················· | | |
| G | 5842 hCoV-19/USA/VA-DCLS-1045/2020\|EPI_ISL_526928\|2020- | ···················· | ···················· | ···················· | ···················· | ···················· | ···················· | | |
| G | 5843 hCoV-19/USA/VA-DCLS-1046/2020\|EPI_ISL_526929\|2020- | ···················· | ···················· | ···················· | ···················· | ···················· | ···················· | | |
| G | 5844 hCoV-19/USA/VA-DCLS-1047/2020\|EPI_ISL_526930\|2020- | ···················· | ···················· | ···················· | ···················· | ···················· | ···················· | | |
| G | 5845 hCoV-19/USA/VA-DCLS-1048/2020\|EPI_ISL_526931\|2020- | ···················· | ···················· | ···················· | ···················· | ···················· | ···················· | | |
| G | 5846 hCoV-19/Colombia/AMA-INS-105161/2020\|EPI_ISL_52693 | ···················· | ···················· | ···················· | ···················· | ···················· | ···················· | | |
| G | 5847 hCoV-19/Colombia/AMA-INS-103045/2020\|EPI_ISL_52693 | ···················· | ···················· | ···················· | ···················· | ···················· | ···················· | | |
| G | 5848 hCoV-19/Faroe Islands/HFS-04/2020\|EPI_ISL_526937\|2 | ···················· | ···················· | ···················· | ···················· | ···················· | ···················· | | |
| G | 5849 hCoV-19/Faroe Islands/HFS-05/2020\|EPI_ISL_526938\|2 | ···················· | ···················· | ···················· | ···················· | ···················· | ···················· | | |
| G | 5850 hCoV-19/Faroe Islands/HFS-06/2020\|EPI_ISL_526939\|2 | ···················· | ···················· | ···················· | ···················· | ···················· | ···················· | | |
| G | 5851 hCoV-19/Faroe Islands/HFS-10/2020\|EPI_ISL_526940\|2 | ···················· | ···················· | ···················· | ···················· | ···················· | ···················· | | |
| G | 5852 hCoV-19/Faroe Islands/HFS-11/2020\|EPI_ISL_526941\|2 | ···················· | ···················· | ···················· | ···················· | ···················· | ···················· | | |
| G | 5853 hCoV-19/Faroe Islands/HFS-30/2020\|EPI_ISL_526943\|2 | ···················· | ···················· | ···················· | ···················· | ···················· | ···················· | | |
| G | 5854 hCoV-19/Switzerland/BS-UHB-42186303/2020\|EPI_ISL_5 | ···················· | ···················· | ···················· | ···················· | ···················· | ···················· | | |
| G | 5855 hCoV-19/Switzerland/BL-UHB-42186366/2020\|EPI_ISL_5 | ···················· | ···················· | ···················· | ···················· | ···················· | ···················· | | |
| G | 5856 hCoV-19/Switzerland/BS-UHB-42186379/2020\|EPI_ISL_5 | ···················· | ···················· | ···················· | ···················· | ···················· | ···················· | | |
| G | 5857 hCoV-19/Switzerland/BS-UHB-42186390/2020\|EPI_ISL_5 | ···················· | ···················· | ···················· | ···················· | ···················· | ···················· | | |
| G | 5858 hCoV-19/Switzerland/BS-UHB-42186394/2020\|EPI_ISL_5 | ···················· | ···················· | ···················· | ···················· | ···················· | ···················· | | |
| G | 5859 hCoV-19/Switzerland/BL-UHB-42186401/2020\|EPI_ISL_5 | ···················· | ···················· | ···················· | ···················· | ···················· | ···················· | | |
| G | 5860 hCoV-19/Switzerland/BS-UHB-42186413/2020\|EPI_ISL_5 | ···················· | ···················· | ···················· | ···················· | ···················· | ···················· | | |
| G | 5861 hCoV-19/Switzerland/BS-UHB-42186439/2020\|EPI_ISL_5 | ···················· | ···················· | ···················· | ···················· | ···················· | ···················· | | |
| G | 5862 hCoV-19/Switzerland/BS-UHB-42186504/2020\|EPI_ISL_5 | ···················· | ···················· | ···················· | ···················· | ···················· | ···················· | | |
| G | 5863 hCoV-19/Switzerland/BS-UHB-42186509/2020\|EPI_ISL_5 | ···················· | ···················· | ···················· | ···················· | ···················· | ···················· | | |
| G | 5864 hCoV-19/Switzerland/BL-UHB-42186528/2020\|EPI_ISL_5 | ···················· | ···················· | ···················· | ···················· | ···················· | ···················· | | |
| G | 5865 hCoV-19/Switzerland/BL-UHB-42186550/2020\|EPI_ISL_5 | ···················· | ···················· | ···················· | ···················· | ···················· | ···················· | | |
| G | 5866 hCoV-19/Switzerland/BL-UHB-42186574/2020\|EPI_ISL_5 | ···················· | ···················· | ···················· | ···················· | ···················· | ···················· | | |
| G | 5867 hCoV-19/Switzerland/BS-UHB-42186648/2020\|EPI_ISL_5 | ···················· | ···················· | ···················· | ···················· | ···················· | ···················· | | |
| G | 5868 hCoV-19/Switzerland/BL-UHB-42186686/2020\|EPI_ISL_5 | ···················· | ···················· | ···················· | ···················· | ···················· | ···················· | | |
| G | 5869 hCoV-19/Switzerland/BS-UHB-42186690/2020\|EPI_ISL_5 | ···················· | ···················· | ···················· | ···················· | ···················· | ···················· | | |
| G | 5870 hCoV-19/Switzerland/BS-UHB-42186718/2020\|EPI_ISL_5 | ···················· | ···················· | ···················· | ···················· | ···················· | ···················· | | |
| G | 5871 hCoV-19/Switzerland/BS-UHB-42187080/2020\|EPI_ISL_5 | ···················· | ···················· | ···················· | ···················· | ···················· | ···················· | | |
| G | 5872 hCoV-19/Switzerland/BS-UHB-42187348/2020\|EPI_ISL_5 | ···················· | ···················· | ···················· | ···················· | ···················· | ···················· | | |
| G | 5873 hCoV-19/Switzerland/BS-UHB-42187354/2020\|EPI_ISL_5 | ···················· | ···················· | ···················· | ···················· | ···················· | ···················· | | |
| G | 5874 hCoV-19/Switzerland/BL-UHB-42187456/2020\|EPI_ISL_5 | ···················· | ···················· | ···················· | ···················· | ···················· | ···················· | | |
| G | 5875 hCoV-19/Switzerland/BS-UHB-42187670/2020\|EPI_ISL_5 | ···················· | ···················· | ···················· | ···················· | ···················· | ···················· | | |
| G | 5876 hCoV-19/Switzerland/BS-UHB-42187831/2020\|EPI_ISL_5 | ···················· | ···················· | ···················· | ···················· | ···················· | ···················· | | |
| G | 5877 hCoV-19/Switzerland/BL-UHB-42187833/2020\|EPI_ISL_5 | ···················· | ···················· | ···················· | ···················· | ···················· | ···················· | | |
| G | 5878 hCoV-19/Switzerland/BS-UHB-42187926/2020\|EPI_ISL_5 | ···················· | ···················· | ···················· | ···················· | ···················· | ···················· | | |
| G | 5879 hCoV-19/Switzerland/BS-UHB-42187936/2020\|EPI_ISL_5 | ···················· | ···················· | ···················· | ···················· | ···················· | ···················· | | |
| G | 5880 hCoV-19/Switzerland/AG-UHB-42187967/2020\|EPI_ISL_5 | ···················· | ···················· | ···················· | ···················· | ···················· | ···················· | | |
| G | 5881 hCoV-19/Switzerland/BS-UHB-42188011/2020\|EPI_ISL_5 | ···················· | ···················· | ···················· | ···················· | ···················· | ···················· | | |
| G | 5882 hCoV-19/Switzerland/BL-UHB-42188088/2020\|EPI_ISL_5 | ···················· | ···················· | ···················· | ···················· | ···················· | ···················· | | |
| G | 5883 hCoV-19/Switzerland/BS-UHB-42188149/2020\|EPI_ISL_5 | ···················· | ···················· | ···················· | ···················· | ···················· | ···················· | | |
| G | 5884 hCoV-19/Switzerland/BS-UHB-42188447/2020\|EPI_ISL_5 | ···················· | ···················· | ···················· | ···················· | ···················· | ···················· | | |
| G | 5885 hCoV-19/Switzerland/BS-UHB-42188679/2020\|EPI_ISL_5 | ···················· | ···················· | ···················· | ···················· | ···················· | ···················· | | |
| G | 5886 hCoV-19/Switzerland/BS-UHB-42188884/2020\|EPI_ISL_5 | ···················· | ···················· | ···················· | ···················· | ···················· | ···················· | | |
| G | 5887 hCoV-19/Switzerland/BL-UHB-42189201/2020\|EPI_ISL_5 | ···················· | ···················· | ···················· | ···················· | ···················· | ···················· | | |
| G | 5888 hCoV-19/Switzerland/BL-UHB-42189725/2020\|EPI_ISL_5 | ···················· | ·A·················· | ···················· | ···················· | ···················· | ···················· | | |
| G | 5892 hCoV-19/England/LIVE-A84B3/2020\|EPI_ISL_517170\|202 | ···················· | ···················· | ···················· | ···················· | ···················· | ···················· | | |
| G | 5893 hCoV-19/England/LIVE-A84C2/2020\|EPI_ISL_517171\|202 | ···················· | ···················· | ···················· | ···················· | ···················· | ···················· | | |
| G | 5894 hCoV-19/England/LIVE-A84D1/2020\|EPI_ISL_517172\|202 | ···················· | ···················· | ···················· | ···················· | ···················· | ···················· | | |
| G | 5895 hCoV-19/England/LIVE-A84FF/2020\|EPI_ISL_517173\|202 | ···················· | ···················· | ···················· | ···················· | ···················· | ···················· | | |
| G | 5897 hCoV-19/England/LIVE-A851A/2020\|EPI_ISL_517175\|202 | ···················· | ···················· | ···················· | ···················· | ···················· | ···················· | | |
| G | 5898 hCoV-19/England/LIVE-A8529/2020\|EPI_ISL_517176\|202 | ···················· | ···················· | ···················· | ···················· | ···················· | ···················· | | |
| G | 5899 hCoV-19/England/LIVE-A8538/2020\|EPI_ISL_517177\|202 | ···················· | ···················· | ···················· | ···················· | ···················· | ···················· | | |
| G | 5900 hCoV-19/England/LIVE-A8547/2020\|EPI_ISL_517178\|202 | ···················· | ···················· | ···················· | ···················· | ···················· | ···················· | | |
| G | 5901 hCoV-19/England/LIVE-A8565/2020\|EPI_ISL_517180\|202 | ···················· | ···················· | ···················· | ···················· | ···················· | ···················· | | |
| G | 5902 hCoV-19/England/LIVE-A8583/2020\|EPI_ISL_517182\|202 | ···················· | ···················· | ···················· | ···················· | ···················· | ···················· | | |
| G | 5903 hCoV-19/England/LIVE-A85A1/2020\|EPI_ISL_517184\|202 | ···················· | ···················· | ···················· | ···················· | ···················· | ···················· | | |
| G | 5904 hCoV-19/England/LIVE-A85B0/2020\|EPI_ISL_517185\|202 | ···················· | ···················· | ···················· | ···················· | ···················· | ···················· | | |
| G | 5906 hCoV-19/England/LIVE-A85DE/2020\|EPI_ISL_517187\|202 | ···················· | ···················· | ···················· | ···················· | ······NN···········N | ···················· | 1 | 1 |
| G | 5907 hCoV-19/England/LIVE-A85ED/2020\|EPI_ISL_517188\|202 | ···················· | ···················· | ···················· | ···················· | ···················· | ···················· | | |
| G | 5908 hCoV-19/England/LIVE-A8608/2020\|EPI_ISL_517189\|202 | ···················· | ···················· | ···················· | ···················· | ···················· | ···················· | | |
| G | 5909 hCoV-19/Australia/VIC5684/2020\|EPI_ISL_518490\|2020 | ···················· | ···················· | ···················· | ···················· | ···················· | ···················· | | |
| G | 5910 hCoV-19/Australia/VIC5685/2020\|EPI_ISL_518491\|2020 | ···················· | ···················· | ···················· | ···················· | ···················· | ···················· | | |
| G | 5911 hCoV-19/Australia/VIC5686/2020\|EPI_ISL_518492\|2020 | ···················· | ···················· | ···················· | ···················· | ···················· | ···················· | | |
| G | 5912 hCoV-19/Australia/VIC5687/2020\|EPI_ISL_518493\|2020 | ···················· | ···················· | ···················· | ···················· | ···················· | ···················· | | |
| G | 5913 hCoV-19/Australia/VIC5688/2020\|EPI_ISL_518494\|2020 | ···················· | ···················· | ···················· | ···················· | ···················· | ···················· | | |
| G | 5914 hCoV-19/Australia/VIC5689/2020\|EPI_ISL_518495\|2020 | ···················· | ···················· | ···················· | ···················· | ···················· | ···················· | | |
| G | 5915 hCoV-19/Australia/VIC5690/2020\|EPI_ISL_518496\|2020 | ···················· | ···················· | ···················· | ···················· | ···················· | ···················· | | |
| G | 5916 hCoV-19/Australia/VIC5691/2020\|EPI_ISL_518497\|2020 | ···················· | ···················· | ···················· | ···················· | ···················· | ···················· | | |
| G | 5917 hCoV-19/Australia/VIC5692/2020\|EPI_ISL_518498\|2020 | ···················· | ···················· | ···················· | ···················· | ···················· | ···················· | | |
| G | 5918 hCoV-19/Australia/VIC5693/2020\|EPI_ISL_518499\|2020 | ···················· | ···················· | ···················· | ···················· | ···················· | ···················· | | |
| G | 5919 hCoV-19/mink/Netherlands/NB-EMC-1-14/2020\|EPI_ISL_ | ···················· | ···················· | ···················· | ···················· | ···················· | ···················· | | |
| G | 5920 hCoV-19/mink/Netherlands/NB-EMC-1-16/2020\|EPI_ISL_ | ···················· | ···················· | ···················· | ···················· | ···················· | ···················· | | |
| G | 5921 hCoV-19/mink/Netherlands/NB-EMC-1-17/2020\|EPI_ISL_ | ···················· | ···················· | ···················· | ···················· | ···················· | ···················· | | |
| G | 5922 hCoV-19/mink/Netherlands/NB-EMC-1-18/2020\|EPI_ISL_ | ···················· | ···················· | ···················· | ···················· | ···················· | ···················· | | |
| G | 5923 hCoV-19/mink/Netherlands/NB-EMC-1-2/2020\|EPI_ISL_5 | ···················· | ···················· | ···················· | ···················· | ···················· | ···················· | | |
| G | 5924 hCoV-19/mink/Netherlands/NB-EMC-12-11/2020\|EPI_ISL | ···················· | ···················· | ···················· | ···················· | ···················· | ···················· | | |
| G | 5925 hCoV-19/mink/Netherlands/NB-EMC-12-13/2020\|EPI_ISL | ···················· | ···················· | ···················· | ···················· | ···················· | ···················· | | |

| | | | | | | | | |
|---|---|---|---|---|---|---|---|---|
| G | 5926 | hCoV-19/mink/Netherlands/NB-EMC-12-23/2020\|EPI_ISL | .................... | .................... | .................... | .................... | .................... | .................... |
| G | 5927 | hCoV-19/mink/Netherlands/NB-EMC-12-37/2020\|EPI_ISL | .................... | .................... | .................... | .................... | .................... | .................... |
| G | 5928 | hCoV-19/mink/Netherlands/NB-EMC-12-7/2020\|EPI_ISL_ | .................... | .................... | .................... | .................... | .................... | .................... |
| G | 5929 | hCoV-19/mink/Netherlands/NB-EMC-1-3/2020\|EPI_ISL_5 | .................... | .................... | .................... | .................... | .................... | .................... |
| G | 5930 | hCoV-19/mink/Netherlands/NB-EMC-13-11/2020\|EPI_ISL | .................... | .................... | .................... | .................... | .................... | .................... |
| G | 5931 | hCoV-19/mink/Netherlands/NB-EMC-13-4/2020\|EPI_ISL_ | .................... | .................... | .................... | .................... | .................... | .................... |
| G | 5932 | hCoV-19/mink/Netherlands/NB-EMC-13-9/2020\|EPI_ISL_ | .................... | .................... | .................... | .................... | .................... | .................... |
| G | 5933 | hCoV-19/mink/Netherlands/NB-EMC-1-4/2020\|EPI_ISL_5 | .................... | .................... | .................... | .................... | .................... | .................... |
| G | 5934 | hCoV-19/mink/Netherlands/NB-EMC-14-21/2020\|EPI_ISL | .................... | .................... | .................... | .................... | .................... | .................... |
| G | 5935 | hCoV-19/mink/Netherlands/NB-EMC-14-25/2020\|EPI_ISL | .................... | .................... | .................... | .................... | .................... | .................... |
| G | 5936 | hCoV-19/mink/Netherlands/NB-EMC-14-3/2020\|EPI_ISL_ | .................... | .................... | .................... | .................... | .................... | .................... |
| G | 5937 | hCoV-19/mink/Netherlands/NB-EMC-14-37/2020\|EPI_ISL | .................... | .................... | .................... | .................... | .................... | .................... |
| G | 5938 | hCoV-19/mink/Netherlands/NB-EMC-15-11/2020\|EPI_ISL | .................... | .................... | .................... | .................... | .................... | .................... |
| G | 5939 | hCoV-19/mink/Netherlands/NB-EMC-15-2/2020\|EPI_ISL_ | .................... | .................... | .................... | .................... | .................... | .................... |
| G | 5940 | hCoV-19/mink/Netherlands/NB-EMC-15-3/2020\|EPI_ISL_ | .................... | .................... | .................... | .................... | .................... | .................... |
| G | 5941 | hCoV-19/mink/Netherlands/NB-EMC-15-7/2020\|EPI_ISL_ | .................... | .................... | .................... | .................... | .................... | .................... |
| G | 5942 | hCoV-19/mink/Netherlands/NB-EMC-15-8/2020\|EPI_ISL_ | .................... | .................... | .................... | .................... | .................... | .................... |
| G | 5943 | hCoV-19/mink/Netherlands/NB-EMC-1-6/2020\|EPI_ISL_5 | .................... | .................... | .................... | .................... | .................... | .................... |
| G | 5944 | hCoV-19/mink/Netherlands/NB-EMC-1-7/2020\|EPI_ISL_5 | .................... | .................... | .................... | .................... | .................... | .................... |
| G | 5945 | hCoV-19/mink/Netherlands/NB-EMC-17-10/2020\|EPI_ISL | .................... | .................... | .................... | .................... | .................... | .................... |
| G | 5946 | hCoV-19/mink/Netherlands/NB-EMC-17-16/2020\|EPI_ISL | .................... | .................... | .................... | .................... | .................... | .................... |
| G | 5947 | hCoV-19/mink/Netherlands/NB-EMC-17-19/2020\|EPI_ISL | .................... | .................... | .................... | .................... | .................... | .................... |
| G | 5948 | hCoV-19/mink/Netherlands/NB-EMC-17-3/2020\|EPI_ISL_ | .................... | .................... | .................... | .................... | .................... | .................... |
| G | 5949 | hCoV-19/mink/Netherlands/NB-EMC-17-6/2020\|EPI_ISL_ | .................... | .................... | .................... | .................... | .................... | .................... |
| G | 5950 | hCoV-19/USA/UT-08542/2020\|EPI_ISL_524303\|2020-05-0 | .................... | .................... | .................... | .................... | .................... | .................... |
| G | 5951 | hCoV-19/USA/UT-08543/2020\|EPI_ISL_524304\|2020-05-2 | .................... | .................... | .................... | .................... | .................... | .................... |
| G | 5952 | hCoV-19/USA/UT-08544/2020\|EPI_ISL_524305\|2020-05-1 | .................... | .A.................. | .................... | .................... | .................... | .................... |
| G | 5953 | hCoV-19/USA/UT-08547/2020\|EPI_ISL_524307\|2020-05-1 | .................... | .................... | .................... | .................... | .................... | .................... |
| G | 5954 | hCoV-19/USA/UT-08549/2020\|EPI_ISL_524309\|2020-05-1 | .................... | .................... | .................... | .................... | .................... | .................... |
| G | 5955 | hCoV-19/USA/UT-08552/2020\|EPI_ISL_524311\|2020-05-1 | .................... | .................... | .................... | .................... | .................... | .................... |
| G | 5956 | hCoV-19/USA/UT-08554/2020\|EPI_ISL_524313\|2020-05-2 | .................... | .................... | .................... | .................... | .................... | .................... |
| G | 5957 | hCoV-19/USA/UT-08555/2020\|EPI_ISL_524314\|2020-05-1 | .................... | .................... | .................... | .................... | .................... | .................... |
| G | 5958 | hCoV-19/USA/UT-08558/2020\|EPI_ISL_524316\|2020-05-1 | .................... | .................... | .................... | .................... | .................... | .................... |
| G | 5959 | hCoV-19/USA/UT-08563/2020\|EPI_ISL_524320\|2020-05-1 | .................... | .................... | .................... | .................... | .................... | .................... |
| G | 5960 | hCoV-19/USA/UT-08564/2020\|EPI_ISL_524321\|2020-05-0 | .................... | .................... | .................... | .................... | .................... | .................... |
| G | 5961 | hCoV-19/USA/UT-08565/2020\|EPI_ISL_524322\|2020-05-2 | .................... | .................... | .................... | .................... | .................... | .................... |
| G | 5962 | hCoV-19/USA/UT-08566/2020\|EPI_ISL_524323\|2020-05-1 | .................... | .................... | .................... | .................... | .................... | .................... |
| G | 5963 | hCoV-19/USA/UT-08567/2020\|EPI_ISL_524324\|2020-05-1 | .................... | .................... | .................... | .................... | .................... | .................... |
| G | 5964 | hCoV-19/USA/UT-08568/2020\|EPI_ISL_524325\|2020-05-1 | .................... | .................... | .................... | .................... | .................... | .................... |
| G | 5965 | hCoV-19/USA/UT-08570/2020\|EPI_ISL_524327\|2020-05-1 | .................... | .................... | .................... | .................... | .................... | .................... |
| G | 5966 | hCoV-19/USA/UT-08579/2020\|EPI_ISL_524333\|2020-05-1 | .................... | .................... | .................... | .................... | .................... | .................... |
| G | 5967 | hCoV-19/USA/UT-08582/2020\|EPI_ISL_524335\|2020-05-2 | .................... | .................... | .................... | .................... | .................... | .................... |
| G | 5968 | hCoV-19/USA/UT-08586/2020\|EPI_ISL_524337\|2020-05-0 | .................... | .................... | .................... | .................... | .................... | .................... |
| G | 5969 | hCoV-19/USA/UT-08594/2020\|EPI_ISL_524341\|2020-05-2 | .................... | .................... | .................... | .................... | .................... | .................... |
| G | 5970 | hCoV-19/USA/UT-08595/2020\|EPI_ISL_524342\|2020-05-2 | .................... | .................... | .................... | .................... | .................... | .................... |
| G | 5971 | hCoV-19/USA/UT-08596/2020\|EPI_ISL_524343\|2020-05-2 | .................... | .................... | .................... | .................... | .................... | .................... |
| G | 5972 | hCoV-19/USA/UT-08597/2020\|EPI_ISL_524344\|2020-05-0 | .................... | .................... | .................... | .................... | .................... | .................... |
| G | 5973 | hCoV-19/USA/UT-08598/2020\|EPI_ISL_524345\|2020-05-2 | .................... | .................... | .................... | .................... | .................... | .................... |
| G | 5974 | hCoV-19/USA/UT-08599/2020\|EPI_ISL_524346\|2020-05-1 | .................... | .................... | .................... | .................... | .................... | .................... |
| G | 5975 | hCoV-19/USA/UT-08601/2020\|EPI_ISL_524348\|2020-05-1 | .................... | .................... | .................... | .................... | .................... | .................... |
| G | 5976 | hCoV-19/USA/UT-08606/2020\|EPI_ISL_524350\|2020-05-2 | .................... | .................... | .................... | .................... | .................... | .................... |
| G | 5977 | hCoV-19/USA/UT-08607/2020\|EPI_ISL_524351\|2020-05-2 | .................... | .................... | .................... | .................... | .................... | .................... |
| G | 5978 | hCoV-19/USA/UT-08608/2020\|EPI_ISL_524352\|2020-05-0 | .................... | .................... | .................... | .................... | .................... | .................... |
| G | 5979 | hCoV-19/USA/UT-08609/2020\|EPI_ISL_524353\|2020-05-2 | .................... | .................... | .................... | .................... | .................... | .................... |
| G | 5980 | hCoV-19/USA/UT-08612/2020\|EPI_ISL_524354\|2020-05-1 | .................... | .................... | .................... | .................... | .................... | .................... |
| G | 5981 | hCoV-19/USA/UT-08614/2020\|EPI_ISL_524355\|2020-05-1 | .................... | .................... | .................... | .................... | .................... | .................... |
| G | 5982 | hCoV-19/USA/UT-08616/2020\|EPI_ISL_524357\|2020-05-2 | .................... | .................... | .................... | .................... | ...........G........ | .................... |
| G | 5983 | hCoV-19/USA/UT-08617/2020\|EPI_ISL_524358\|2020-05-2 | .................... | .................... | .................... | .................... | .................... | .................... |
| G | 5984 | hCoV-19/USA/UT-08619/2020\|EPI_ISL_524359\|2020-05-0 | .................... | .................... | .................... | .................... | .................... | .................... |
| G | 5985 | hCoV-19/USA/UT-08620/2020\|EPI_ISL_524360\|2020-05-2 | .................... | .................... | .................... | .................... | .................... | .................... |
| G | 5986 | hCoV-19/USA/UT-08622/2020\|EPI_ISL_524362\|2020-05-2 | .................... | .................... | .................... | .................... | .................... | .................... |
| G | 5987 | hCoV-19/USA/UT-08627/2020\|EPI_ISL_524365\|2020-05-2 | .................... | .................... | .................... | .................... | .................... | .................... |
| G | 5988 | hCoV-19/USA/UT-08629/2020\|EPI_ISL_524367\|2020-05-1 | .................... | .................... | .................... | .................... | .................... | .................... |
| G | 5989 | hCoV-19/USA/UT-08630/2020\|EPI_ISL_524368\|2020-05-1 | .................... | .................... | .................... | .................... | .................... | .................... |
| G | 5990 | hCoV-19/USA/NY-Wadsworth-26730-01/2020\|EPI_ISL_525 | .................... | .................... | .................... | .................... | .................... | .................... |
| G | 5991 | hCoV-19/USA/NY-Wadsworth-27452-01/2020\|EPI_ISL_525 | .................... | .................... | .................... | .................... | .................... | .................... |
| G | 5992 | hCoV-19/USA/NY-Wadsworth-27593-01/2020\|EPI_ISL_525 | .................... | .................... | .................... | .................... | .................... | .................... |
| G | 5993 | hCoV-19/USA/NY-Wadsworth-27938-01/2020\|EPI_ISL_525 | .................... | .................... | .................... | .................... | .................... | .................... |
| G | 5994 | hCoV-19/USA/NY-Wadsworth-28157-01/2020\|EPI_ISL_525 | .................... | .................... | .................... | .................... | .................... | .................... |
| G | 5995 | hCoV-19/USA/NY-Wadsworth-29389-01/2020\|EPI_ISL_525 | .................... | .................... | .................... | .................... | .................... | .................... |
| G | 5996 | hCoV-19/USA/NY-Wadsworth-30593-01/2020\|EPI_ISL_525 | .................... | .................... | .................... | .................... | .................... | .................... |
| G | 5997 | hCoV-19/USA/NY-Wadsworth-30673-01/2020\|EPI_ISL_525 | .................... | .................... | .................... | .................... | .................... | .................... |
| G | 5998 | hCoV-19/USA/NY-Wadsworth-30826-01/2020\|EPI_ISL_525 | .................... | .................... | .................... | .................... | .................... | .................... |
| G | 5999 | hCoV-19/USA/NY-Wadsworth-31058-01/2020\|EPI_ISL_525 | .................... | .................... | .................... | .................... | .................... | .................... |
| G | 6000 | hCoV-19/USA/NY-Wadsworth-31562-01/2020\|EPI_ISL_525 | .................... | .................... | .................... | .................... | .................... | .................... |
| G | 6001 | hCoV-19/USA/NY-Wadsworth-32689-01/2020\|EPI_ISL_525 | .................... | .................... | .................... | .................... | .................... | .................... |
| G | 6002 | hCoV-19/USA/NY-Wadsworth-33279-01/2020\|EPI_ISL_525 | .................... | .................... | .................... | .................... | .................... | .................... |
| G | 6003 | hCoV-19/USA/NY-Wadsworth-33281-01/2020\|EPI_ISL_525 | .................... | .................... | .................... | .................... | .................... | .................... |
| G | 6004 | hCoV-19/USA/NY-Wadsworth-33282-01/2020\|EPI_ISL_525 | .................... | .................... | .................... | .................... | .................... | .................... |
| G | 6005 | hCoV-19/USA/NY-Wadsworth-33283-01/2020\|EPI_ISL_525 | .................... | .................... | .................... | .................... | .................... | .................... |

| | | | | | | | | |
|---|---|---|---|---|---|---|---|---|
| G | 6006 | hCoV-19/USA/NY-Wadsworth-33284-01/2020\|EPI_ISL_525 | .................... | .................... | .................... | .................... | .................... | .................... |
| G | 6007 | hCoV-19/USA/NY-Wadsworth-34682-01/2020\|EPI_ISL_525 | .................... | .................... | .................... | .................... | .................... | .................... |
| G | 6008 | hCoV-19/USA/NY-Wadsworth-34728-01/2020\|EPI_ISL_525 | .................... | .................... | .................... | .................... | .................... | .................... |
| G | 6009 | hCoV-19/USA/NY-Wadsworth-34804-01/2020\|EPI_ISL_525 | .................... | .................... | .................... | .................... | .................... | .................... |
| G | 6010 | hCoV-19/USA/NY-Wadsworth-34848-01/2020\|EPI_ISL_525 | .................... | .................... | .................... | .................... | .................... | .................... |
| G | 6011 | hCoV-19/USA/NY-Wadsworth-34855-01/2020\|EPI_ISL_525 | .................... | .................... | .................... | .................... | .................... | .................... |
| G | 6012 | hCoV-19/USA/NY-Wadsworth-36877-01/2020\|EPI_ISL_525 | .................... | .................... | .................... | .................... | .................... | .................... |
| G | 6013 | hCoV-19/USA/NY-Wadsworth-36878-01/2020\|EPI_ISL_525 | .................... | .................... | .................... | .................... | .................... | .................... |
| G | 6014 | hCoV-19/USA/NY-Wadsworth-36879-01/2020\|EPI_ISL_525 | .................... | .................... | .................... | .................... | .................... | .................... |
| G | 6015 | hCoV-19/USA/NY-Wadsworth-37630-01/2020\|EPI_ISL_525 | .................... | .................... | .................... | .................... | .................... | .................... |
| G | 6016 | hCoV-19/USA/NY-Wadsworth-37640-01/2020\|EPI_ISL_525 | .................... | .................... | .................... | .................... | .................... | .................... |
| G | 6017 | hCoV-19/USA/NY-Wadsworth-38698-01/2020\|EPI_ISL_525 | .................... | .................... | .................... | .................... | .................... | .................... |
| G | 6018 | hCoV-19/USA/NY-Wadsworth-39179-01/2020\|EPI_ISL_525 | .................... | .................... | .................... | .................... | .................... | .................... |
| G | 6019 | hCoV-19/USA/NY-Wadsworth-39537-01/2020\|EPI_ISL_525 | .................... | .................... | .................... | .................... | .................... | .................... |
| G | 6020 | hCoV-19/USA/NY-Wadsworth-39698-01/2020\|EPI_ISL_525 | .................... | .................... | .................... | .................... | .................... | .................... |
| G | 6021 | hCoV-19/USA/NY-Wadsworth-40198-01/2020\|EPI_ISL_525 | .................... | .................... | .................... | .................... | .................... | .................... |
| G | 6022 | hCoV-19/USA/NY-Wadsworth-41104-01/2020\|EPI_ISL_525 | .................... | .................... | .................... | .................... | .................... | .................... |
| G | 6023 | hCoV-19/USA/NY-Wadsworth-41107-01/2020\|EPI_ISL_525 | .................... | .................... | .................... | .................... | .................... | .................... |
| G | 6024 | hCoV-19/USA/NY-Wadsworth-41904-01/2020\|EPI_ISL_525 | .................... | .................... | .................... | .................... | .................... | .................... |
| G | 6025 | hCoV-19/USA/NY-Wadsworth-42409-01/2020\|EPI_ISL_525 | .................... | .................... | .................... | .................... | .................... | .................... |
| G | 6026 | hCoV-19/USA/NY-Wadsworth-43319-01/2020\|EPI_ISL_525 | .................... | .................... | .................... | .................... | .................... | .................... |
| G | 6027 | hCoV-19/USA/NY-Wadsworth-43604-01/2020\|EPI_ISL_525 | .................... | .................... | .................... | .................... | .................... | .................... |
| G | 6028 | hCoV-19/USA/NY-Wadsworth-44440-01/2020\|EPI_ISL_525 | .................... | .................... | .................... | .................... | .................... | .................... |
| G | 6029 | hCoV-19/USA/NY-Wadsworth-44556-01/2020\|EPI_ISL_525 | .................... | .................... | .................... | .................... | .................... | .................... |
| G | 6030 | hCoV-19/USA/NY-Wadsworth-44557-01/2020\|EPI_ISL_525 | .................... | .................... | .................... | .................... | .................... | .................... |
| G | 6031 | hCoV-19/USA/NY-Wadsworth-44600-01/2020\|EPI_ISL_525 | .................... | .................... | .................... | .................... | .................... | .................... |
| G | 6032 | hCoV-19/USA/NY-Wadsworth-44639-01/2020\|EPI_ISL_525 | .................... | .................... | .................... | .................... | .................... | .................... |
| G | 6033 | hCoV-19/USA/NY-Wadsworth-44644-01/2020\|EPI_ISL_525 | .................... | .................... | .................... | .................... | .................... | .................... |
| G | 6034 | hCoV-19/USA/NY-Wadsworth-45057-01/2020\|EPI_ISL_525 | .................... | .................... | .................... | .................... | .................... | .................... |
| G | 6035 | hCoV-19/USA/NY-Wadsworth-45936-01/2020\|EPI_ISL_525 | .................... | .................... | .................... | .................... | .................... | .................... |
| G | 6036 | hCoV-19/USA/NY-Wadsworth-46188-01/2020\|EPI_ISL_525 | .................... | .................... | .................... | .................... | .................... | .................... |
| G | 6037 | hCoV-19/USA/NY-Wadsworth-46314-01/2020\|EPI_ISL_525 | .................... | .................... | .................... | .................... | .................... | .................... |
| G | 6038 | hCoV-19/USA/NY-Wadsworth-46315-01/2020\|EPI_ISL_525 | .................... | .................... | .................... | .................... | .................... | .................... |
| G | 6039 | hCoV-19/USA/NY-WC-46318-01/2020\|EPI_ISL_525678\|202 | .................... | .................... | .................... | .................... | .................... | .................... |
| G | 6040 | hCoV-19/USA/NY-Wadsworth-46319-01/2020\|EPI_ISL_525 | .................... | .................... | .................... | .................... | .................... | .................... |
| G | 6041 | hCoV-19/USA/NY-Wadsworth-46320-01/2020\|EPI_ISL_525 | .................... | .................... | .................... | .................... | .................... | .................... |
| G | 6042 | hCoV-19/USA/NY-Wadsworth-50101-01/2020\|EPI_ISL_525 | .................... | .................... | .................... | .................... | .................... | .................... |
| G | 6043 | hCoV-19/USA/NY-Wadsworth-50104-01/2020\|EPI_ISL_525 | .................... | .................... | .................... | .................... | .................... | .................... |
| G | 6044 | hCoV-19/USA/NY-Wadsworth-50112-01/2020\|EPI_ISL_525 | .................... | .................... | .................... | .................... | .................... | .................... |
| G | 6045 | hCoV-19/USA/NY-Wadsworth-50113-01/2020\|EPI_ISL_525 | .................... | .................... | .................... | .................... | .................... | .................... |
| G | 6046 | hCoV-19/USA/NY-Wadsworth-51085-01/2020\|EPI_ISL_525 | .................... | .................... | .................... | .................... | .................... | .................... |
| G | 6047 | hCoV-19/USA/NY-Wadsworth-57118-01/2020\|EPI_ISL_525 | .................... | .................... | .................... | .................... | .................... | .................... |
| G | 6048 | hCoV-19/Colombia/INS-CUN-AG2/2020\|EPI_ISL_526958\|2 | .................... | .................... | .................... | .................... | .................... | .................... |
| G | 6050 | hCoV-19/Colombia/TOL-INS-0345-T20/2020\|EPI_ISL_526 | .................... | .................... | .................... | .................... | .................... | .................... |
| G | 6051 | hCoV-19/Colombia/TOL-INS-0341-T15/2020\|EPI_ISL_526 | .................... | .................... | .................... | .................... | .................... | .................... |
| G | 6052 | hCoV-19/Colombia/TOL-INS-0330-T5/2020\|EPI_ISL_5269 | .................... | .................... | .................... | .................... | .................... | .................... |
| G | 6053 | hCoV-19/Colombia/DC-INS-289/2020\|EPI_ISL_526967\|20 | .................... | .................... | .................... | .................... | .................... | .................... |
| G | 6055 | hCoV-19/Egypt/EGY-020/2020\|EPI_ISL_526975\|2020-08- | .................... | .................... | .................... | .................... | .................... | .................... |
| G | 6056 | hCoV-19/Egypt/EGY-021/2020\|EPI_ISL_526976\|2020-08- | .................... | .................... | .................... | .................... | .................... | .................... |
| G | 6057 | hCoV-19/Egypt/EGY-022/2020\|EPI_ISL_526977\|2020-08- | .................... | .................... | .................... | .................... | .................... | .................... |
| G | 6058 | hCoV-19/Egypt/EGY-023/2020\|EPI_ISL_526978\|2020-08- | .................... | .................... | .................... | .................... | .................... | .................... |
| G | 6059 | hCoV-19/Egypt/EGY-024/2020\|EPI_ISL_526979\|2020-08- | .................... | .................... | .................... | .................... | .................... | .................... |
| G | 6060 | hCoV-19/Egypt/EGY-025/2020\|EPI_ISL_526980\|2020-08- | .................... | .................... | .................... | .................... | .................... | .................... |
| G | 6061 | hCoV-19/Egypt/EGY-026/2020\|EPI_ISL_526981\|2020-08- | .................... | .................... | .................... | .................... | .................... | .................... |
| G | 6062 | hCoV-19/Egypt/EGY-027/2020\|EPI_ISL_526982\|2020-08- | .................... | .................... | .................... | .................... | .................... | .................... |
| G | 6063 | hCoV-19/Egypt/EGY-028/2020\|EPI_ISL_526983\|2020-08- | .................... | .................... | .................... | .................... | .................... | .................... |
| G | 6064 | hCoV-19/Egypt/EGY-029/2020\|EPI_ISL_526984\|2020-08- | .................... | .................... | .................... | .................... | .................... | .................... |
| G | 6065 | hCoV-19/Egypt/EGY-030/2020\|EPI_ISL_526985\|2020-08- | .................... | .................... | .................... | .................... | .................... | .................... |
| G | 6066 | hCoV-19/Egypt/EGY-S031/2020\|EPI_ISL_526986\|2020-08 | .................... | .................... | .................... | .................... | .................... | .................... |
| G | 6067 | hCoV-19/Egypt/EGY-S032/2020\|EPI_ISL_526987\|2020-08 | .................... | .................... | .................... | .................... | .................... | .................... |
| G | 6068 | hCoV-19/Egypt/EGY-S033/2020\|EPI_ISL_526988\|2020-08 | .................... | .................... | .................... | .................... | .................... | .................... |
| G | 6069 | hCoV-19/Egypt/EGY-S034/2020\|EPI_ISL_526989\|2020-08 | .................... | .................... | .................... | .................... | .................... | .................... |
| G | 6070 | hCoV-19/Egypt/EGY-S035/2020\|EPI_ISL_526990\|2020-08 | .................... | .................... | .................... | .................... | .................... | .................... |
| G | 6071 | hCoV-19/Egypt/EGY-S036/2020\|EPI_ISL_526991\|2020-08 | .................... | .................... | .................... | .................... | .................... | .................... |
| G | 6072 | hCoV-19/Egypt/EGY-S037/2020\|EPI_ISL_526992\|2020-08 | .................... | .................... | .................... | .................... | .................... | .................... |
| G | 6073 | hCoV-19/Egypt/EGY-S038/2020\|EPI_ISL_526993\|2020-08 | .................... | .................... | .................... | .................... | .................... | .................... |
| G | 6074 | hCoV-19/Egypt/EGY-S039/2020\|EPI_ISL_526994\|2020-08 | .................... | .................... | .................... | .................... | .................... | .................... |
| G | 6075 | hCoV-19/Egypt/EGY-S040/2020\|EPI_ISL_526995\|2020-08 | .................... | .................... | .................... | .................... | .................... | .................... |
| G | 6076 | hCoV-19/Egypt/EGY-S030/2020\|EPI_ISL_526996\|2020-08 | .................... | .................... | .................... | .................... | .................... | .................... |
| G | 6077 | hCoV-19/Egypt/EGY-041/2020\|EPI_ISL_526997\|2020-08- | .................... | .................... | .................... | .................... | .................... | .................... |
| G | 6078 | hCoV-19/Egypt/EGY-042/2020\|EPI_ISL_526998\|2020-08- | .................... | .................... | .................... | .................... | .................... | .................... |
| G | 6079 | hCoV-19/Egypt/EGY-044/2020\|EPI_ISL_526999\|2020-08- | .................... | .................... | .................... | .................... | .................... | .................... |
| G | 6080 | hCoV-19/USA/WA-S2768/2020\|EPI_ISL_530125\|2020-05-1 | .................... | .................... | .................... | .................... | .................... | .................... |
| G | 6081 | hCoV-19/USA/WA-S2769/2020\|EPI_ISL_530126\|2020-07-1 | .................... | .................... | .................... | .................... | .................... | .................... |
| G | 6082 | hCoV-19/USA/WA-S2770/2020\|EPI_ISL_530127\|2020-08-0 | .................... | .................... | .................... | .................... | .................... | .................... |
| G | 6083 | hCoV-19/USA/WA-S2771/2020\|EPI_ISL_530128\|2020-08-0 | .................... | .................... | .................... | .................... | .................... | .................... |
| G | 6084 | hCoV-19/USA/WA-S2772/2020\|EPI_ISL_530129\|2020-08-0 | .................... | .A.................. | .................... | .................... | .................... | .................... |
| G | 6085 | hCoV-19/USA/WA-S2774/2020\|EPI_ISL_530131\|2020-08-0 | .................... | .................... | .................... | .................... | .................... | .................... |
| G | 6086 | hCoV-19/USA/WA-S2775/2020\|EPI_ISL_530132\|2020-08-0 | .................... | .A.................. | .................... | .................... | .................... | .................... |
| G | 6087 | hCoV-19/USA/WA-S2779/2020\|EPI_ISL_530136\|2020-08-0 | .................... | .................... | .................... | .................... | .................... | .................... |

G 6088 hCoV-19/USA/WA-S2780/2020|EPI_ISL_530137|2020-08-1 ···················· ·A·················· ···················· ···················· ···················· ····················
G 6089 hCoV-19/USA/WA-S2781/2020|EPI_ISL_530138|2020-08-1 ···················· ·A·················· ···················· ···················· ···················· ····················
G 6090 hCoV-19/USA/WA-S2782/2020|EPI_ISL_530139|2020-08-1 ···················· ···A················ ···················· ···················· ···················· ····················
G 6091 hCoV-19/USA/WA-S2783/2020|EPI_ISL_530140|2020-08-1 ···················· ·A·················· ···················· ···················· ···················· ····················
G 6092 hCoV-19/USA/WA-S2784/2020|EPI_ISL_530141|2020-08-1 ···················· ···················· ···················· ···················· ···················· ····················
G 6093 hCoV-19/USA/WA-S2785/2020|EPI_ISL_530142|2020-08-1 ···················· ···················· ···················· ···················· ···················· ····················
G 6094 hCoV-19/USA/WA-S2786/2020|EPI_ISL_530143|2020-08-1 ···················· ···················· ···················· ···················· ···················· ····················
G 6095 hCoV-19/USA/WA-S2787/2020|EPI_ISL_530144|2020-08-1 ···················· ···················· ···················· ···················· ···················· ····················
G 6096 hCoV-19/USA/WA-S2788/2020|EPI_ISL_530145|2020-08-1 ···················· ···················· ···················· ···················· ···················· ····················
G 6097 hCoV-19/USA/WA-S2789/2020|EPI_ISL_530146|2020-08-1 ···················· ···················· ···················· ···················· ···················· ····················
G 6098 hCoV-19/USA/WA-S2790/2020|EPI_ISL_530147|2020-08-1 ···················· ···················· ···················· ···················· ···················· ····················
G 6099 hCoV-19/USA/WA-S2791/2020|EPI_ISL_530148|2020-08-1 ···················· ···················· ···················· ···················· ···················· ····················
G 6100 hCoV-19/USA/WA-S2792/2020|EPI_ISL_530149|2020-08-1 ···················· ···················· ···················· ···················· ···················· ····················
G 6101 hCoV-19/USA/WA-S2793/2020|EPI_ISL_530150|2020-08-1 ···················· ···················· ···················· ···················· ···················· ····················
G 6102 hCoV-19/USA/WA-S2794/2020|EPI_ISL_530151|2020-08-1 ···················· ·A·················· ···················· ···················· ···················· ····················
G 6103 hCoV-19/USA/WA-S2795/2020|EPI_ISL_530152|2020-08-1 ···················· ···················· ···················· ···················· ···················· ····················
G 6104 hCoV-19/USA/WA-S2797/2020|EPI_ISL_530154|2020-08-1 ···················· ·A·················· ···················· ···················· ···················· ····················
G 6105 hCoV-19/USA/WA-S2798/2020|EPI_ISL_530155|2020-08-2 ···················· ·A·················· ···················· ···················· ···················· ····················
G 6106 hCoV-19/USA/WA-S2799/2020|EPI_ISL_530156|2020-08-2 ···················· ·A·················· ···················· ···················· ···················· ····················
G 6107 hCoV-19/USA/WA-S2800/2020|EPI_ISL_530157|2020-08-2 ···················· ·A·················· ···················· ···················· ···················· ····················
G 6108 hCoV-19/USA/WA-S2801/2020|EPI_ISL_530158|2020-08-2 ···················· ·A·················· ···················· ···················· ···················· ····················
G 6109 hCoV-19/USA/WA-S2802/2020|EPI_ISL_530159|2020-08-2 ···················· ···················· ···················· ···················· ···················· ····················
G 6110 hCoV-19/USA/WA-S2804/2020|EPI_ISL_530161|2020-08-2 ···················· ·A·················· ···················· ···················· ···················· ··A·················
G 6111 hCoV-19/USA/WA-S2805/2020|EPI_ISL_530162|2020-08-2 ···················· ·A·················· ···················· ···················· ···················· ··A·················
G 6112 hCoV-19/USA/MN-MDH-1567/2020|EPI_ISL_530163|2020-0 ···················· ···················· ···················· ···················· ···················· ····················
G 6113 hCoV-19/USA/MN-MDH-1568/2020|EPI_ISL_530164|2020-0 ···················· ···················· ···················· ···················· ···················· ····················
G 6114 hCoV-19/USA/MN-MDH-1569/2020|EPI_ISL_530165|2020-0 ···················· ···················· ···················· ···················· ···················· ····················
G 6115 hCoV-19/USA/MN-MDH-1570/2020|EPI_ISL_530166|2020-0 ···················· ···················· ···················· ···················· ···················· ····················
G 6116 hCoV-19/USA/MN-MDH-1571/2020|EPI_ISL_530167|2020-0 ···················· ·A·················· ···················· ···················· ···················· ····················
G 6117 hCoV-19/USA/MN-MDH-1572/2020|EPI_ISL_530168|2020-0 ···················· ·A·················· ···················· ···················· ···················· ····················
G 6118 hCoV-19/USA/MN-MDH-1573/2020|EPI_ISL_530169|2020-0 ···················· ·A·················· ···················· ···················· ···················· ····················
G 6119 hCoV-19/USA/MN-MDH-1574/2020|EPI_ISL_530170|2020-0 ···················· ···················· ···················· ···················· ···················· ····················
G 6120 hCoV-19/USA/MN-MDH-1575/2020|EPI_ISL_530171|2020-0 ···················· ···················· ···················· ···················· ···················· ····················
G 6121 hCoV-19/USA/MN-MDH-1576/2020|EPI_ISL_530172|2020-0 ···················· ···················· ···················· ···················· ···················· ····················
G 6122 hCoV-19/USA/MN-MDH-1577/2020|EPI_ISL_530173|2020-0 ···················· ···················· ···················· ···················· ···················· ····················
G 6123 hCoV-19/USA/MN-MDH-1578/2020|EPI_ISL_530174|2020-0 ···················· ···················· ···················· ···················· ···················· ····················
G 6124 hCoV-19/USA/MN-MDH-1579/2020|EPI_ISL_530175|2020-0 ···················· ···················· ···················· ···················· ···················· ····················
G 6125 hCoV-19/USA/MN-MDH-1580/2020|EPI_ISL_530176|2020-0 ···················· ···················· ···················· ···················· ···················· ····················
G 6126 hCoV-19/USA/MN-MDH-1581/2020|EPI_ISL_530177|2020-0 ···················· ···················· ···················· ···················· ···················· ····················
G 6127 hCoV-19/USA/MN-MDH-1582/2020|EPI_ISL_530178|2020-0 ···················· ···················· ···················· ···················· ···················· ····················
G 6128 hCoV-19/USA/MN-MDH-1583/2020|EPI_ISL_530179|2020-0 ···················· ···················· ···················· ···················· ···················· ····················
G 6129 hCoV-19/USA/MN-MDH-1584/2020|EPI_ISL_530180|2020-0 ···················· ···················· ···················· ···················· ···················· ····················
G 6130 hCoV-19/USA/MN-MDH-1585/2020|EPI_ISL_530181|2020-0 ···················· ···················· ···················· ···················· ···················· ····················
G 6131 hCoV-19/USA/MN-MDH-1586/2020|EPI_ISL_530182|2020-0 ···················· ···················· ···················· ···················· ···················· ····················
G 6132 hCoV-19/USA/MN-MDH-1587/2020|EPI_ISL_530183|2020-0 ···················· ···················· ···················· ···················· ···················· ····················
G 6133 hCoV-19/USA/MN-MDH-1588/2020|EPI_ISL_530184|2020-0 ···················· ···················· ···················· ···················· ···················· ····················
G 6134 hCoV-19/USA/MN-MDH-1589/2020|EPI_ISL_530185|2020-0 ···················· ···················· ···················· ···················· ···················· ····················
G 6135 hCoV-19/USA/MN-MDH-1590/2020|EPI_ISL_530186|2020-0 ···················· ···················· ···················· ···················· ···················· ····················
G 6136 hCoV-19/USA/MN-MDH-1591/2020|EPI_ISL_530187|2020-0 ···················· ···················· ···················· ···················· ···················· ····················
G 6137 hCoV-19/USA/MN-MDH-1592/2020|EPI_ISL_530188|2020-0 ···················· ···················· ···················· ···················· ···················· ····················
G 6138 hCoV-19/USA/MN-MDH-1593/2020|EPI_ISL_530189|2020-0 ···················· ···················· ···················· ···················· ···················· ····················
G 6139 hCoV-19/USA/MN-MDH-1594/2020|EPI_ISL_530190|2020-0 ···················· ···················· ···················· ···················· ···················· ····················
G 6140 hCoV-19/USA/MN-MDH-1595/2020|EPI_ISL_530191|2020-0 ···················· ···················· ···················· ···················· ···················· ····················
G 6141 hCoV-19/USA/MN-MDH-1596/2020|EPI_ISL_530192|2020-0 ···················· ···················· ···················· ···················· ···················· ····················
G 6142 hCoV-19/USA/MN-MDH-1597/2020|EPI_ISL_530193|2020-0 ···················· ···················· ···················· ···················· ···················· ····················
G 6143 hCoV-19/USA/MN-MDH-1598/2020|EPI_ISL_530194|2020-0 ···················· ···················· ···················· ···················· ···················· ····················
G 6144 hCoV-19/USA/MN-MDH-1599/2020|EPI_ISL_530195|2020-0 ···················· ···················· ···················· ···················· ···················· ····················
G 6145 hCoV-19/England/LIVE-A918E/2020|EPI_ISL_517270|202 ···················· ···················· ···················· ···················· ···················· ····················
G 6147 hCoV-19/England/LIVE-A91AC/2020|EPI_ISL_517272|202 ···················· ···················· ···················· ···················· ···················· ····················
G 6148 hCoV-19/England/LIVE-A91BB/2020|EPI_ISL_517273|202 ···················· ···················· ···················· ···················· ···················· ····················
G 6149 hCoV-19/England/LIVE-A91E8/2020|EPI_ISL_517275|202 ···················· ···················· ···················· ···················· ···················· ····················
G 6150 hCoV-19/England/LIVE-A91F7/2020|EPI_ISL_517276|202 ···················· ···················· ···················· ···················· ···················· ····················
G 6151 hCoV-19/England/LIVE-A9212/2020|EPI_ISL_517278|202 ···················· ···················· ···················· ···················· ···················· ····················
G 6152 hCoV-19/England/LIVE-A9221/2020|EPI_ISL_517279|202 ···················· ···················· ···················· ···················· ···················· ····················
G 6153 hCoV-19/Australia/VIC5694/2020|EPI_ISL_518500|2020 ···················· ···················· ···················· ···················· ···················· ····················
G 6154 hCoV-19/Australia/VIC5696/2020|EPI_ISL_518501|2020 ···················· ···················· ···················· ···················· ···················· ····················
G 6155 hCoV-19/Australia/VIC5697/2020|EPI_ISL_518502|2020 ···················· ···················· ···················· ···················· ···················· ····················
G 6156 hCoV-19/Australia/VIC5698/2020|EPI_ISL_518503|2020 ···················· ···················· ···················· ···················· ···················· ····················
G 6157 hCoV-19/Australia/VIC5700/2020|EPI_ISL_518504|2020 ···················· ···················· ···················· ···················· ···················· ····················
G 6158 hCoV-19/Australia/VIC5701/2020|EPI_ISL_518505|2020 ···················· ···················· ···················· ···················· ···················· ····················
G 6159 hCoV-19/Australia/VIC5702/2020|EPI_ISL_518506|2020 ···················· ···················· ···················· ···················· ···················· ····················
G 6160 hCoV-19/Australia/VIC5703/2020|EPI_ISL_518507|2020 ···················· ···················· ···················· ···················· ···················· ····················
G 6161 hCoV-19/Australia/VIC5704/2020|EPI_ISL_518508|2020 ···················· ···················· ···················· ···················· ···················· ····················
G 6162 hCoV-19/Australia/VIC5706/2020|EPI_ISL_518509|2020 ···················· ···················· ···················· ···················· ···················· ····················
G 6163 hCoV-19/Australia/VIC5707/2020|EPI_ISL_518510|2020 ···················· ···················· ···················· ···················· ···················· ····················
G 6164 hCoV-19/Australia/VIC5708/2020|EPI_ISL_518511|2020 ···················· ···················· ···················· ···················· ···················· ····················
G 6165 hCoV-19/Australia/VIC5711/2020|EPI_ISL_518514|2020 ···················· ···················· ···················· ···················· ···················· ····················
G 6166 hCoV-19/Australia/VIC5712/2020|EPI_ISL_518515|2020 ···················· ···················· ···················· ···················· ···················· ····················
G 6167 hCoV-19/Australia/VIC5713/2020|EPI_ISL_518516|2020 ···················· ···················· ···················· ···················· ···················· ····················
G 6168 hCoV-19/Australia/VIC5714/2020|EPI_ISL_518517|2020 ···················· ···················· ···················· ···················· ···················· ····················

| | | | | | | | | |
|---|---|---|---|---|---|---|---|---|
| G | 6169 | hCoV-19/Australia/VIC5716/2020\|EPI_ISL_518518\|2020 | ···················· | ···················· | ····················· | ···················· | ···················· | ···················· |
| G | 6171 | hCoV-19/Australia/VIC5718/2020\|EPI_ISL_518520\|2020 | ···················· | ···················· | ····················· | ···················· | ···················· | ···················· |
| G | 6172 | hCoV-19/Australia/VIC5720/2020\|EPI_ISL_518522\|2020 | ···················· | ···················· | ····················· | ···················· | ···················· | ···················· |
| G | 6173 | hCoV-19/Australia/VIC5721/2020\|EPI_ISL_518523\|2020 | ···················· | ···················· | ····················· | ···················· | ···················· | ···················· |
| G | 6174 | hCoV-19/Australia/VIC5723/2020\|EPI_ISL_518524\|2020 | ···················· | ···················· | ····················· | ···················· | ···················· | ···················· |
| G | 6175 | hCoV-19/Australia/VIC5724/2020\|EPI_ISL_518525\|2020 | ···················· | ···················· | ····················· | ···················· | ···················· | ···················· |
| G | 6176 | hCoV-19/Australia/VIC5725/2020\|EPI_ISL_518526\|2020 | ···················· | ···················· | ····················· | ···················· | ···················· | ···················· |
| G | 6177 | hCoV-19/Australia/VIC5726/2020\|EPI_ISL_518527\|2020 | ···················· | ···················· | ····················· | ···················· | ···················· | ···················· |
| G | 6178 | hCoV-19/Australia/VIC5727/2020\|EPI_ISL_518528\|2020 | ···················· | ···················· | ····················· | ···················· | ···················· | ···················· |
| G | 6179 | hCoV-19/Australia/VIC5728/2020\|EPI_ISL_518529\|2020 | ···················· | ···················· | ····················· | ···················· | ···················· | ···················· |
| G | 6180 | hCoV-19/Australia/VIC4996/2020\|EPI_ISL_519800\|2020 | ···················· | ···················· | ····················· | ···················· | ···················· | ···················· |
| G | 6181 | hCoV-19/Australia/VIC4997/2020\|EPI_ISL_519801\|2020 | ···················· | ···················· | ····················· | ···················· | ···················· | ···················· |
| G | 6182 | hCoV-19/Australia/VIC4998/2020\|EPI_ISL_519802\|2020 | ···················· | ···················· | ····················· | ···················· | ···················· | ···················· |
| G | 6183 | hCoV-19/Australia/VIC4999/2020\|EPI_ISL_519803\|2020 | ···················· | ···················· | ····················· | ···················· | ···················· | ···················· |
| G | 6184 | hCoV-19/Australia/VIC5000/2020\|EPI_ISL_519804\|2020 | ···················· | ···················· | ····················· | ···················· | ···················· | ···················· |
| G | 6185 | hCoV-19/Australia/VIC5001/2020\|EPI_ISL_519805\|2020 | ···················· | ···················· | ····················· | ···················· | ···················· | ···················· |
| G | 6186 | hCoV-19/Australia/VIC5002/2020\|EPI_ISL_519806\|2020 | ···················· | ···················· | ····················· | ···················· | ···················· | ···················· |
| G | 6187 | hCoV-19/Australia/VIC5003/2020\|EPI_ISL_519807\|2020 | ···················· | ···················· | ····················· | ···················· | ···················· | ···················· |
| G | 6188 | hCoV-19/Australia/VIC5004/2020\|EPI_ISL_519808\|2020 | ···················· | ···················· | ····················· | ···················· | ···················· | ···················· |
| G | 6189 | hCoV-19/Australia/VIC5005/2020\|EPI_ISL_519809\|2020 | ···················· | ···················· | ····················· | ···················· | ···················· | ···················· |
| G | 6190 | hCoV-19/Australia/VIC5006/2020\|EPI_ISL_519810\|2020 | ···················· | ···················· | ····················· | ···················· | ···················· | ···················· |
| G | 6191 | hCoV-19/Australia/VIC5007/2020\|EPI_ISL_519811\|2020 | ···················· | ···················· | ····················· | ···················· | ···················· | ···················· |
| G | 6192 | hCoV-19/Australia/VIC5008/2020\|EPI_ISL_519812\|2020 | ···················· | ···················· | ····················· | ···················· | ···················· | ···················· |
| G | 6193 | hCoV-19/Australia/VIC5009/2020\|EPI_ISL_519813\|2020 | ···················· | ···················· | ····················· | ···················· | ···················· | ···················· |
| G | 6194 | hCoV-19/Australia/VIC5010/2020\|EPI_ISL_519814\|2020 | ···················· | ···················· | ····················· | ···················· | ···················· | ···················· |
| G | 6195 | hCoV-19/Australia/VIC5011/2020\|EPI_ISL_519815\|2020 | ···················· | ···················· | ····················· | ···················· | ···················· | ···················· |
| G | 6196 | hCoV-19/Australia/VIC5012/2020\|EPI_ISL_519816\|2020 | ···················· | ···················· | ····················· | ···················· | ···················· | ···················· |
| G | 6197 | hCoV-19/Australia/VIC5014/2020\|EPI_ISL_519818\|2020 | ···················· | ···················· | ····················· | ···················· | ···················· | ···················· |
| G | 6198 | hCoV-19/Australia/VIC5015/2020\|EPI_ISL_519819\|2020 | ···················· | ···················· | ····················· | ···················· | ···················· | ···················· |
| G | 6199 | hCoV-19/Australia/VIC5016/2020\|EPI_ISL_519820\|2020 | ···················· | ···················· | ····················· | ···················· | ···················· | ···················· |
| G | 6200 | hCoV-19/Australia/VIC5017/2020\|EPI_ISL_519821\|2020 | ···················· | ···················· | ····················· | ···················· | ···················· | ···················· |
| G | 6201 | hCoV-19/Australia/VIC5018/2020\|EPI_ISL_519822\|2020 | ···················· | ···················· | ····················· | ···················· | ···················· | ···················· |
| G | 6202 | hCoV-19/Australia/VIC5019/2020\|EPI_ISL_519823\|2020 | ···················· | ···················· | ····················· | ···················· | ···················· | ···················· |
| G | 6203 | hCoV-19/Australia/VIC5020/2020\|EPI_ISL_519824\|2020 | ···················· | ···················· | ····················· | ···················· | ···················· | ···················· |
| G | 6204 | hCoV-19/Australia/VIC5021/2020\|EPI_ISL_519825\|2020 | ···················· | ···················· | ····················· | ···················· | ···················· | ···················· |
| G | 6205 | hCoV-19/Australia/VIC5022/2020\|EPI_ISL_519826\|2020 | ···················· | ···················· | ····················· | ···················· | ···················· | ···················· |
| G | 6206 | hCoV-19/Australia/VIC5023/2020\|EPI_ISL_519827\|2020 | ···················· | ···················· | ····················· | ···················· | ···················· | ···················· |
| G | 6207 | hCoV-19/Australia/VIC5024/2020\|EPI_ISL_519828\|2020 | ···················· | ···················· | ····················· | ···················· | ···················· | ···················· |
| G | 6208 | hCoV-19/Australia/VIC5025/2020\|EPI_ISL_519829\|2020 | ···················· | ···················· | ····················· | ···················· | ···················· | ···················· |
| G | 6209 | hCoV-19/Australia/VIC5026/2020\|EPI_ISL_519830\|2020 | ···················· | ···················· | ····················· | ···················· | ···················· | ···················· |
| G | 6210 | hCoV-19/Australia/VIC5027/2020\|EPI_ISL_519831\|2020 | ···················· | ···················· | ····················· | ···················· | ···················· | ···················· |
| G | 6211 | hCoV-19/Australia/VIC5028/2020\|EPI_ISL_519832\|2020 | ···················· | ···················· | ····················· | ···················· | ···················· | ···················· |
| G | 6212 | hCoV-19/Australia/VIC5029/2020\|EPI_ISL_519833\|2020 | ···················· | ···················· | ····················· | ···················· | ···················· | ···················· |
| G | 6213 | hCoV-19/Australia/VIC5030/2020\|EPI_ISL_519834\|2020 | ···················· | ···················· | ····················· | ···················· | ···················· | ···················· |
| G | 6214 | hCoV-19/Australia/VIC5031/2020\|EPI_ISL_519835\|2020 | ···················· | ···················· | ····················· | ···················· | ···················· | ···················· |
| G | 6215 | hCoV-19/Australia/VIC5032/2020\|EPI_ISL_519836\|2020 | ···················· | ···················· | ····················· | ···················· | ···················· | ···················· |
| G | 6216 | hCoV-19/Australia/VIC5033/2020\|EPI_ISL_519837\|2020 | ···················· | ···················· | ····················· | ···················· | ···················· | ···················· |
| G | 6217 | hCoV-19/Australia/VIC5034/2020\|EPI_ISL_519838\|2020 | ···················· | ···················· | ····················· | ···················· | ···················· | ···················· |
| G | 6218 | hCoV-19/Australia/VIC5035/2020\|EPI_ISL_519839\|2020 | ···················· | ···················· | ····················· | ···················· | ···················· | ···················· |
| G | 6219 | hCoV-19/Australia/VIC5036/2020\|EPI_ISL_519840\|2020 | ···················· | ···················· | ····················· | ···················· | ···················· | ···················· |
| G | 6220 | hCoV-19/Australia/VIC5037/2020\|EPI_ISL_519841\|2020 | ···················· | ···················· | ····················· | ···················· | ···················· | ···················· |
| G | 6221 | hCoV-19/Australia/VIC5038/2020\|EPI_ISL_519842\|2020 | ···················· | ···················· | ····················· | ···················· | ···················· | ···················· |
| G | 6222 | hCoV-19/Australia/VIC5039/2020\|EPI_ISL_519843\|2020 | ···················· | ···················· | ····················· | ···················· | ···················· | ···················· |
| G | 6223 | hCoV-19/Australia/VIC5040/2020\|EPI_ISL_519844\|2020 | ···················· | ···················· | ····················· | ···················· | ···················· | ···················· |
| G | 6224 | hCoV-19/Australia/VIC5041/2020\|EPI_ISL_519845\|2020 | ···················· | ···················· | ····················· | ···················· | ···················· | ···················· |
| G | 6225 | hCoV-19/Australia/VIC5042/2020\|EPI_ISL_519846\|2020 | ···················· | ···················· | ····················· | ···················· | ···················· | ···················· |
| G | 6226 | hCoV-19/Australia/VIC5043/2020\|EPI_ISL_519847\|2020 | ···················· | ···················· | ····················· | ···················· | ···················· | ···················· |
| G | 6227 | hCoV-19/Australia/VIC5044/2020\|EPI_ISL_519848\|2020 | ···················· | ···················· | ····················· | ···················· | ···················· | ···················· |
| G | 6228 | hCoV-19/Australia/VIC5045/2020\|EPI_ISL_519849\|2020 | ···················· | ···················· | ····················· | ···················· | ···················· | ···················· |
| G | 6229 | hCoV-19/Australia/VIC5046/2020\|EPI_ISL_519850\|2020 | ···················· | ···················· | ····················· | ···················· | ···················· | ···················· |
| G | 6230 | hCoV-19/Australia/VIC5047/2020\|EPI_ISL_519851\|2020 | ···················· | ···················· | ····················· | ···················· | ···················· | ···················· |
| G | 6231 | hCoV-19/Australia/VIC5048/2020\|EPI_ISL_519852\|2020 | ···················· | ···················· | ····················· | ···················· | ···················· | ···················· |
| G | 6232 | hCoV-19/Australia/VIC5050/2020\|EPI_ISL_519853\|2020 | ···················· | ···················· | ····················· | ···················· | ···················· | ···················· |
| G | 6233 | hCoV-19/Australia/VIC5051/2020\|EPI_ISL_519854\|2020 | ···················· | ···················· | ····················· | ···················· | ···················· | ···················· |
| G | 6234 | hCoV-19/Australia/VIC5052/2020\|EPI_ISL_519855\|2020 | ···················· | ···················· | ····················· | ···················· | ···················· | ···················· |
| G | 6235 | hCoV-19/Australia/VIC5053/2020\|EPI_ISL_519856\|2020 | ···················· | ···················· | ····················· | ···················· | ···················· | ···················· |
| G | 6236 | hCoV-19/Australia/VIC5054/2020\|EPI_ISL_519857\|2020 | ···················· | ···················· | ····················· | ···················· | ···················· | ···················· |
| G | 6237 | hCoV-19/Australia/VIC5055/2020\|EPI_ISL_519858\|2020 | ···················· | ···················· | ····················· | ···················· | ···················· | ···················· |
| G | 6238 | hCoV-19/Australia/VIC5056/2020\|EPI_ISL_519859\|2020 | ···················· | ···················· | ····················· | ···················· | ···················· | ···················· |
| G | 6239 | hCoV-19/Australia/VIC3093/2020\|EPI_ISL_520800\|2020 | ···················· | ···················· | ····················· | ···················· | ···················· | ···················· |
| G | 6240 | hCoV-19/Australia/VIC3094/2020\|EPI_ISL_520801\|2020 | ···················· | ···················· | ····················· | ···················· | ···················· | ···················· |
| G | 6243 | hCoV-19/Australia/VIC3097/2020\|EPI_ISL_520804\|2020 | ···················· | ···················· | ····················· | ···················· | ···················· | ···················· |
| G | 6244 | hCoV-19/Australia/VIC3098/2020\|EPI_ISL_520805\|2020 | ···················· | ···················· | ····················· | ···················· | ···················· | ···················· |
| G | 6245 | hCoV-19/Australia/VIC3099/2020\|EPI_ISL_520806\|2020 | ···················· | ···················· | ····················· | ···················· | ···················· | ···················· |
| G | 6246 | hCoV-19/Australia/VIC3100/2020\|EPI_ISL_520807\|2020 | ···················· | ···················· | ····················· | ···················· | ···················· | ···················· |
| G | 6247 | hCoV-19/Australia/VIC3101/2020\|EPI_ISL_520808\|2020 | ···················· | ···················· | ····················· | ···················· | ···················· | ···················· |
| G | 6248 | hCoV-19/Australia/VIC3102/2020\|EPI_ISL_520809\|2020 | ···················· | ···················· | ····················· | ···················· | ···················· | ···················· |
| G | 6250 | hCoV-19/Australia/VIC3105/2020\|EPI_ISL_520812\|2020 | ···················· | ···················· | ····················· | ···················· | ···················· | ···················· |
| G | 6251 | hCoV-19/Australia/VIC3106/2020\|EPI_ISL_520813\|2020 | ···················· | ···················· | ····················· | ···················· | ···················· | ···················· |
| G | 6252 | hCoV-19/Australia/VIC3107/2020\|EPI_ISL_520814\|2020 | ···················· | ···················· | ····················· | ···················· | ···················· | ···················· |

| | | | | | | | | |
|---|---|---|---|---|---|---|---|---|
| G | 6253 | hCoV-19/Australia/VIC3108/2020\|EPI_ISL_520815\|2020 | ···················· | ···················· | ···················· | ···················· | ···················· | ···················· |
| G | 6254 | hCoV-19/Australia/VIC3109/2020\|EPI_ISL_520816\|2020 | ···················· | ···················· | ···················· | ···················· | ···················· | ···················· |
| G | 6255 | hCoV-19/Australia/VIC3111/2020\|EPI_ISL_520818\|2020 | ···················· | ···················· | ···················· | ···················· | ···················· | ···················· |
| G | 6256 | hCoV-19/Australia/VIC3112/2020\|EPI_ISL_520819\|2020 | ···················· | ···················· | ···················· | ···················· | ···················· | ···················· |
| G | 6257 | hCoV-19/Australia/VIC3113/2020\|EPI_ISL_520820\|2020 | ···················· | ···················· | ···················· | ···················· | ···················· | ···················· |
| G | 6258 | hCoV-19/Australia/VIC3114/2020\|EPI_ISL_520821\|2020 | ···················· | ···················· | ···················· | ···················· | ···················· | ···················· |
| G | 6259 | hCoV-19/Australia/VIC3115/2020\|EPI_ISL_520822\|2020 | ···················· | ···················· | ···················· | ···················· | ···················· | ···················· |
| G | 6260 | hCoV-19/Australia/VIC3116/2020\|EPI_ISL_520823\|2020 | ···················· | ···················· | ···················· | ···················· | ···················· | ···················· |
| G | 6261 | hCoV-19/Australia/VIC3117/2020\|EPI_ISL_520824\|2020 | ···················· | ···················· | ···················· | ···················· | ···················· | ···················· |
| G | 6262 | hCoV-19/Australia/VIC3118/2020\|EPI_ISL_520825\|2020 | ···················· | ···················· | ···················· | ···················· | ···················· | ···················· |
| G | 6263 | hCoV-19/Australia/VIC3119/2020\|EPI_ISL_520826\|2020 | ···················· | ···················· | ···················· | ···················· | ···················· | ···················· |
| G | 6264 | hCoV-19/Australia/VIC3120/2020\|EPI_ISL_520827\|2020 | ···················· | ···················· | ···················· | ···················· | ···················· | ···················· |
| G | 6265 | hCoV-19/Australia/VIC3121/2020\|EPI_ISL_520828\|2020 | ···················· | ···················· | ···················· | ···················· | ···················· | ···················· |
| G | 6266 | hCoV-19/Australia/VIC3122/2020\|EPI_ISL_520829\|2020 | ···················· | ···················· | ···················· | ···················· | ···················· | ···················· |
| G | 6267 | hCoV-19/Australia/VIC3123/2020\|EPI_ISL_520830\|2020 | ···················· | ···················· | ···················· | ···················· | ···················· | ···················· |
| G | 6268 | hCoV-19/Australia/VIC3124/2020\|EPI_ISL_520831\|2020 | ···················· | ···················· | ···················· | ···················· | ···················· | ···················· |
| G | 6270 | hCoV-19/Australia/VIC3126/2020\|EPI_ISL_520833\|2020 | ···················· | ···················· | ···················· | ···················· | ···················· | ···················· |
| G | 6271 | hCoV-19/Australia/VIC3127/2020\|EPI_ISL_520834\|2020 | ···················· | ···················· | ···················· | ···················· | ···················· | ···················· |
| G | 6272 | hCoV-19/Australia/VIC3128/2020\|EPI_ISL_520835\|2020 | ···················· | ···················· | ···················· | ···················· | ···················· | ···················· |
| G | 6273 | hCoV-19/Australia/VIC3129/2020\|EPI_ISL_520836\|2020 | ···················· | ···················· | ···················· | ···················· | ···················· | ···················· |
| G | 6274 | hCoV-19/Australia/VIC3130/2020\|EPI_ISL_520837\|2020 | ···················· | ···················· | ···················· | ···················· | ···················· | ···················· |
| G | 6275 | hCoV-19/Australia/VIC3131/2020\|EPI_ISL_520838\|2020 | ···················· | ···················· | ···················· | ···················· | ···················· | ···················· |
| G | 6277 | hCoV-19/mink/Netherlands/NB-EMC-18-14/2020\|EPI_ISL | ···················· | ···················· | ···················· | ···················· | ···················· | ···················· |
| G | 6278 | hCoV-19/mink/Netherlands/NB-EMC-18-15/2020\|EPI_ISL | ···················· | ···················· | ···················· | ···················· | ···················· | ···················· |
| G | 6279 | hCoV-19/mink/Netherlands/NB-EMC-18-17/2020\|EPI_ISL | ···················· | ···················· | ···················· | ···················· | ···················· | ···················· |
| G | 6280 | hCoV-19/mink/Netherlands/NB-EMC-18-20/2020\|EPI_ISL | ···················· | ···················· | ···················· | ···················· | ···················· | ···················· |
| G | 6281 | hCoV-19/mink/Netherlands/NB-EMC-18-8/2020\|EPI_ISL_ | ···················· | ···················· | ···················· | ···················· | ···················· | ···················· |
| G | 6282 | hCoV-19/mink/Netherlands/NB-EMC-1-9/2020\|EPI_ISL_5 | ···················· | ···················· | ···················· | ···················· | ···················· | ···················· |
| G | 6283 | hCoV-19/mink/Netherlands/NB-EMC-19-12/2020\|EPI_ISL | ···················· | ···················· | ···················· | ···················· | ···················· | ···················· |
| G | 6284 | hCoV-19/mink/Netherlands/NB-EMC-19-15/2020\|EPI_ISL | ···················· | ···················· | ···················· | ···················· | ···················· | ···················· |
| G | 6285 | hCoV-19/mink/Netherlands/NB-EMC-19-19/2020\|EPI_ISL | ···················· | ···················· | ···················· | ···················· | ···················· | ···················· |
| G | 6286 | hCoV-19/mink/Netherlands/NB-EMC-19-9/2020\|EPI_ISL_ | ···················· | ···················· | ···················· | ···················· | ···················· | ···················· |
| G | 6287 | hCoV-19/mink/Netherlands/NB-EMC-20-1/2020\|EPI_ISL_ | ···················· | ···················· | ···················· | ···················· | ···················· | ···················· |
| G | 6288 | hCoV-19/mink/Netherlands/NB-EMC-20-2/2020\|EPI_ISL_ | ···················· | ···················· | ···················· | ···················· | ···················· | ···················· |
| G | 6289 | hCoV-19/mink/Netherlands/NB-EMC-20-21/2020\|EPI_ISL | ···················· | ···················· | ···················· | ···················· | ···················· | ···················· |
| G | 6290 | hCoV-19/mink/Netherlands/NB-EMC-20-23/2020\|EPI_ISL | ···················· | ···················· | ···················· | ···················· | ···················· | ···················· |
| G | 6291 | hCoV-19/mink/Netherlands/NB-EMC-20-27/2020\|EPI_ISL | ···················· | ···················· | ···················· | ···················· | ···················· | ···················· |
| G | 6292 | hCoV-19/mink/Netherlands/NB-EMC-20-28/2020\|EPI_ISL | ···················· | ···················· | ···················· | ···················· | ···················· | ···················· |
| G | 6293 | hCoV-19/mink/Netherlands/NB-EMC-20-3/2020\|EPI_ISL_ | ···················· | ···················· | ···················· | ···················· | ···················· | ···················· |
| G | 6294 | hCoV-19/mink/Netherlands/NB-EMC-20-34/2020\|EPI_ISL | ···················· | ···················· | ···················· | ···················· | ···················· | ···················· |
| G | 6295 | hCoV-19/mink/Netherlands/NB-EMC-20-4/2020\|EPI_ISL_ | ···················· | ···················· | ···················· | ···················· | ···················· | ···················· |
| G | 6296 | hCoV-19/mink/Netherlands/NB-EMC-20-5/2020\|EPI_ISL_ | ···················· | ···················· | ···················· | ···················· | ···················· | ···················· |
| G | 6297 | hCoV-19/mink/Netherlands/NB-EMC-21-07/2020\|EPI_ISL | ···················· | ···················· | ···················· | ···················· | ······T············· | ···················· |
| G | 6298 | hCoV-19/mink/Netherlands/NB-EMC-21-1/2020\|EPI_ISL_ | ···················· | ···················· | ···················· | ···················· | ······T············· | ···················· |
| G | 6299 | hCoV-19/mink/Netherlands/NB-EMC-21-14/2020\|EPI_ISL | ···················· | ···················· | ···················· | ···················· | ······T············· | ···················· |
| G | 6300 | hCoV-19/mink/Netherlands/NB-EMC-21-19/2020\|EPI_ISL | ···················· | ···················· | ···················· | ···················· | ······T············· | ···················· |
| G | 6301 | hCoV-19/mink/Netherlands/NB-EMC-21-2/2020\|EPI_ISL_ | ···················· | ···················· | ···················· | ···················· | ······T············· | ···················· |
| G | 6302 | hCoV-19/mink/Netherlands/NB-EMC-21-20/2020\|EPI_ISL | ···················· | ···················· | ···················· | ···················· | ······T············· | ···················· |
| G | 6303 | hCoV-19/mink/Netherlands/NB-EMC-21-4/2020\|EPI_ISL_ | ···················· | ···················· | ···················· | ···················· | ······T············· | ···················· |
| G | 6304 | hCoV-19/mink/Netherlands/NB-EMC-2-14/2020\|EPI_ISL_ | ···················· | ···················· | ···················· | ···················· | ····A··············· | ···················· |
| G | 6305 | hCoV-19/mink/Netherlands/NB-EMC-21-5/2020\|EPI_ISL_ | ···················· | ···················· | ···················· | ···················· | ······T············· | ···················· |
| G | 6306 | hCoV-19/mink/Netherlands/NB-EMC-22-1/2020\|EPI_ISL_ | ···················· | ···················· | ···················· | ···················· | ···················· | ···················· |
| G | 6307 | hCoV-19/mink/Netherlands/NB-EMC-2-3/2020\|EPI_ISL_5 | ···················· | ···················· | ···················· | ···················· | ···················· | ···················· |
| G | 6308 | hCoV-19/mink/Netherlands/NB-EMC-23-1/2020\|EPI_ISL_ | ···················· | ···················· | ···················· | ···················· | ···················· | ···················· |
| G | 6309 | hCoV-19/mink/Netherlands/NB-EMC-23-2/2020\|EPI_ISL_ | ···················· | ···················· | ···················· | ···················· | ···················· | ···················· |
| G | 6310 | hCoV-19/mink/Netherlands/NB-EMC-23-4/2020\|EPI_ISL_ | ···················· | ···················· | ···················· | ···················· | ···················· | ···················· |
| G | 6311 | hCoV-19/mink/Netherlands/NB-EMC-23-5/2020\|EPI_ISL_ | ···················· | ···················· | ···················· | ···················· | ···················· | ···················· |
| G | 6312 | hCoV-19/mink/Netherlands/NB-EMC-23-6/2020\|EPI_ISL_ | ···················· | ···················· | ···················· | ···················· | ···················· | ···················· |
| G | 6313 | hCoV-19/mink/Netherlands/NB-EMC-23-7/2020\|EPI_ISL_ | ···················· | ···················· | ···················· | ···················· | ···················· | ···················· |
| G | 6314 | hCoV-19/mink/Netherlands/NB-EMC-25-11/2020\|EPI_ISL | ···················· | ···················· | ···················· | ···················· | ···················· | ···················· |
| G | 6316 | hCoV-19/mink/Netherlands/NB-EMC-2-6/2020\|EPI_ISL_5 | ···················· | ···················· | ···················· | ···················· | ····A··············· | ···················· |
| G | 6317 | hCoV-19/mink/Netherlands/NB-EMC-26-1/2020\|EPI_ISL_ | ···················· | ···················· | ···················· | ···················· | ···················· | ···················· |
| G | 6318 | hCoV-19/mink/Netherlands/NB-EMC-26-2/2020\|EPI_ISL_ | ···················· | ···················· | ···················· | ···················· | ···················· | ···················· |
| G | 6319 | hCoV-19/mink/Netherlands/NB-EMC-3-2/2020\|EPI_ISL_5 | ···················· | ···················· | ···················· | ···················· | ···················· | ···················· |
| G | 6320 | hCoV-19/mink/Netherlands/NB-EMC-3-3/2020\|EPI_ISL_5 | ···················· | ···················· | ···················· | ···················· | ···················· | ···················· |
| G | 6321 | hCoV-19/mink/Netherlands/NB-EMC-3-4/2020\|EPI_ISL_5 | ···················· | ···················· | ···················· | ···················· | ···················· | ···················· |
| G | 6322 | hCoV-19/mink/Netherlands/NB-EMC-3-5/2020\|EPI_ISL_5 | ···················· | ···················· | ···················· | ···················· | ···················· | ···················· |
| G | 6323 | hCoV-19/mink/Netherlands/NB-EMC-3-6/2020\|EPI_ISL_5 | ···················· | ···················· | ···················· | ···················· | ···················· | ···················· |
| G | 6324 | hCoV-19/mink/Netherlands/NB-EMC-4-2/2020\|EPI_ISL_5 | ···················· | ···················· | ···················· | ···················· | ···················· | ···················· |
| G | 6325 | hCoV-19/mink/Netherlands/NB-EMC-5-13/2020\|EPI_ISL_ | ···················· | ···················· | ···················· | ···················· | ···················· | ···················· |
| G | 6326 | hCoV-19/mink/Netherlands/NB-EMC-6-13/2020\|EPI_ISL_ | ···················· | ···················· | ···················· | ···················· | ···················· | ···················· |
| G | 6327 | hCoV-19/mink/Netherlands/NB-EMC-6-14/2020\|EPI_ISL_ | ···················· | ···················· | ···················· | ···················· | ···················· | ···················· |
| G | 6328 | hCoV-19/USA/UT-08632/2020\|EPI_ISL_524370\|2020-05-2 | ···················· | ···················· | ···················· | ···················· | ···················· | ···················· |
| G | 6329 | hCoV-19/USA/UT-08633/2020\|EPI_ISL_524371\|2020-05-2 | ···················· | ···················· | ···················· | ···················· | ···················· | ···················· |
| G | 6330 | hCoV-19/USA/UT-08634/2020\|EPI_ISL_524372\|2020-05-1 | ···················· | ···················· | ···················· | ···················· | ···················· | ···················· |
| G | 6331 | hCoV-19/USA/UT-08635/2020\|EPI_ISL_524373\|2020-05-0 | ···················· | ···················· | ···················· | ···················· | ···················· | ···················· |
| G | 6332 | hCoV-19/USA/UT-08636/2020\|EPI_ISL_524374\|2020-05-0 | ···················· | ···················· | ···················· | ···················· | ···················· | ···················· |
| G | 6333 | hCoV-19/USA/UT-08638/2020\|EPI_ISL_524375\|2020-05-0 | ···················· | ···················· | ···················· | ···················· | ···················· | ···················· |
| G | 6334 | hCoV-19/USA/UT-08639/2020\|EPI_ISL_524376\|2020-05-0 | ···················· | ···················· | ···················· | ···················· | ···················· | ···················· |
| G | 6335 | hCoV-19/USA/UT-08642/2020\|EPI_ISL_524377\|2020-05-2 | ···················· | ···················· | ···················· | ···················· | ···················· | ···················· |

| | | | | | | | | |
|---|---|---|---|---|---|---|---|---|
| G | 6336 hCoV-19/USA/UT-08643/2020\|EPI_ISL_524378\|2020-05-2 | ···················· | ···················· | ···················· | ···················· | ···················· | ···················· | |
| G | 6337 hCoV-19/USA/UT-08649/2020\|EPI_ISL_524381\|2020-05-0 | ···················· | ···················· | ···················· | ···················· | ···················· | ···················· | |
| G | 6338 hCoV-19/USA/UT-08650/2020\|EPI_ISL_524382\|2020-05-0 | ···················· | ···················· | ···················· | ···················· | ···················· | ···················· | |
| G | 6339 hCoV-19/USA/UT-08652/2020\|EPI_ISL_524384\|2020-05-1 | ···················· | ···················· | ···················· | ···················· | ···················· | ···················· | |
| G | 6340 hCoV-19/USA/UT-08656/2020\|EPI_ISL_524385\|2020-05-0 | ···················· | ···················· | ···················· | ···················· | ···················· | ···················· | |
| G | 6341 hCoV-19/USA/UT-08661/2020\|EPI_ISL_524387\|2020-05-2 | ···················· | ···················· | ···················· | ···················· | ···················· | ···················· | |
| G | 6342 hCoV-19/USA/UT-08662/2020\|EPI_ISL_524388\|2020-05-0 | ···················· | ···················· | ···················· | ···················· | ···················· | ···················· | |
| G | 6343 hCoV-19/USA/UT-08663/2020\|EPI_ISL_524389\|2020-05-0 | ···················· | ···················· | ···················· | ···················· | ···················· | ···················· | |
| G | 6344 hCoV-19/USA/UT-08666/2020\|EPI_ISL_524392\|2020-05-0 | ···················· | ···················· | ···················· | ···················· | ···················· | ···················· | |
| G | 6345 hCoV-19/USA/UT-08667/2020\|EPI_ISL_524393\|2020-05-0 | ···················· | ···················· | ···················· | ···················· | ···················· | ···················· | |
| G | 6346 hCoV-19/USA/UT-08669/2020\|EPI_ISL_524394\|2020-05-2 | ···················· | ···················· | ···················· | ···················· | ···················· | ···················· | |
| G | 6347 hCoV-19/USA/UT-08671/2020\|EPI_ISL_524396\|2020-05-0 | ···················· | ···················· | ···················· | ···················· | ···················· | ···················· | |
| G | 6348 hCoV-19/USA/UT-08674/2020\|EPI_ISL_524398\|2020-05-0 | ···················· | ···················· | ···················· | ···················· | ·········A·········· | ···················· | |
| G | 6349 hCoV-19/USA/NY-Wadsworth-17364-01/2020\|EPI_ISL_525 | ···················· | ···················· | ···················· | ···················· | ···················· | ···················· | |
| G | 6350 hCoV-19/USA/NY-Wadsworth-17549-01/2020\|EPI_ISL_525 | ···················· | ···················· | ···················· | ···················· | ···················· | ···················· | |
| G | 6351 hCoV-19/USA/NY-Wadsworth-18631-01/2020\|EPI_ISL_525 | ···················· | ···················· | ···················· | ···················· | ···················· | ···················· | |
| G | 6352 hCoV-19/USA/NY-Wadsworth-18637-01/2020\|EPI_ISL_525 | ···················· | ···················· | ···················· | ···················· | ···················· | ···················· | |
| G | 6353 hCoV-19/USA/NY-Wadsworth-19679-01/2020\|EPI_ISL_525 | ···················· | ···················· | ···················· | ···················· | ···················· | ···················· | |
| G | 6354 hCoV-19/USA/NY-Wadsworth-21196-01/2020\|EPI_ISL_525 | ···················· | ···················· | ···················· | ···················· | ···················· | ···················· | |
| G | 6355 hCoV-19/USA/NY-Wadsworth-21493-01/2020\|EPI_ISL_525 | ···················· | ···················· | ···················· | ···················· | ···················· | ···················· | |
| G | 6356 hCoV-19/USA/NY-Wadsworth-22513-01/2020\|EPI_ISL_525 | ···················· | ···················· | ···················· | ···················· | ···················· | ···················· | |
| G | 6357 hCoV-19/USA/NY-Wadsworth-24149-01/2020\|EPI_ISL_525 | ···················· | ···················· | ···················· | ···················· | ···················· | ···················· | |
| G | 6358 hCoV-19/USA/NY-Wadsworth-24516-01/2020\|EPI_ISL_525 | ···················· | ···················· | ···················· | ···················· | ···················· | ···················· | |
| G | 6359 hCoV-19/USA/NY-Wadsworth-25458-01/2020\|EPI_ISL_525 | ···················· | ···················· | ···················· | ···················· | ···················· | ···················· | |
| G | 6360 hCoV-19/Faroe Islands/HFS-43/2020\|EPI_ISL_526946\|2 | ···················· | ···················· | ···················· | ···················· | ···················· | ···················· | |
| G | 6361 hCoV-19/Colombia/AMA-INS-109363/2020\|EPI_ISL_52694 | ···················· | ···················· | ···················· | ···················· | ···················· | ···················· | |
| G | 6363 hCoV-19/Colombia/AMA-INS-103039-2/2020\|EPI_ISL_526 | ···················· | ···················· | ···················· | ···················· | ···················· | ···················· | |
| G | 6364 hCoV-19/Colombia/NAR-INS-103303-2/2020\|EPI_ISL_526 | ···················· | ···················· | ···················· | ···················· | ···················· | ···················· | |
| G | 6366 hCoV-19/England/NORW-EE76D/2020\|EPI_ISL_529244\|202 | ···················· | ···················· | ···················· | ···················· | ···················· | ···················· | |
| G | 6367 hCoV-19/England/NORW-EE78B/2020\|EPI_ISL_529246\|202 | ···················· | ···················· | ···················· | ···················· | ···················· | ···················· | |
| G | 6368 hCoV-19/England/NORW-EEA19/2020\|EPI_ISL_529247\|202 | ···················· | ···················· | ···················· | ···················· | ···················· | ···················· | |
| G | 6369 hCoV-19/England/NORW-EEB9E/2020\|EPI_ISL_529250\|202 | ···················· | ···················· | ···················· | ···················· | ···················· | ···················· | |
| G | 6370 hCoV-19/England/BIRM-637D9/2020\|EPI_ISL_529251\|202 | ···················· | ···················· | ···················· | ···················· | ···················· | ···················· | |
| G | 6371 hCoV-19/England/BIRM-637E8/2020\|EPI_ISL_529252\|202 | ···················· | ···················· | ···················· | ···················· | ···················· | ···················· | |
| G | 6372 hCoV-19/Wales/PHWC-169E37/2020\|EPI_ISL_529253\|2020 | ···················· | ···················· | ···················· | ···················· | ···················· | ···················· | |
| G | 6373 hCoV-19/USA/MN-MDH-1600/2020\|EPI_ISL_530196\|2020-0 | ···················· | ···················· | ···················· | ···················· | ···················· | ···················· | |
| G | 6374 hCoV-19/USA/MN-MDH-1601/2020\|EPI_ISL_530197\|2020-0 | ···················· | ···················· | ···················· | ···················· | ···················· | ···················· | |
| G | 6375 hCoV-19/USA/MN-MDH-1602/2020\|EPI_ISL_530198\|2020-0 | ···················· | ···················· | ···················· | ···················· | ···················· | ···················· | |
| G | 6376 hCoV-19/USA/MN-MDH-1603/2020\|EPI_ISL_530199\|2020-0 | ···················· | ···················· | ···················· | ···················· | ···················· | ···················· | |
| G | 6377 hCoV-19/USA/MN-MDH-1604/2020\|EPI_ISL_530200\|2020-0 | ···················· | ···················· | ···················· | ···················· | ···················· | ···················· | |
| G | 6378 hCoV-19/USA/MN-MDH-1605/2020\|EPI_ISL_530201\|2020-0 | ···················· | ···················· | ···················· | ···················· | ···················· | ···················· | |
| G | 6379 hCoV-19/USA/MN-MDH-1606/2020\|EPI_ISL_530202\|2020-0 | ···················· | ···················· | ···················· | ···················· | ···················· | ···················· | |
| G | 6380 hCoV-19/USA/MN-MDH-1607/2020\|EPI_ISL_530203\|2020-0 | ···················· | ···················· | ···················· | ···················· | ···················· | ···················· | |
| G | 6381 hCoV-19/USA/MN-MDH-1608/2020\|EPI_ISL_530204\|2020-0 | ···················· | ···················· | ···················· | ···················· | ···················· | ···················· | |
| G | 6382 hCoV-19/USA/MN-MDH-1609/2020\|EPI_ISL_530205\|2020-0 | ···················· | ···················· | ···················· | ···················· | ···················· | ···················· | |
| G | 6383 hCoV-19/USA/MN-MDH-1610/2020\|EPI_ISL_530206\|2020-0 | ···················· | ···················· | ···················· | ···················· | ···················· | ···················· | |
| G | 6384 hCoV-19/USA/MN-MDH-1611/2020\|EPI_ISL_530207\|2020-0 | ···················· | ···················· | ···················· | ···················· | ···················· | ···················· | |
| G | 6385 hCoV-19/USA/MN-MDH-1612/2020\|EPI_ISL_530208\|2020-0 | ···················· | ···················· | ···················· | ···················· | ···················· | ···················· | |
| G | 6386 hCoV-19/USA/MN-MDH-1613/2020\|EPI_ISL_530209\|2020-0 | ···················· | ···················· | ···················· | ···················· | ···················· | ···················· | |
| G | 6387 hCoV-19/USA/MN-MDH-1614/2020\|EPI_ISL_530210\|2020-0 | ···················· | ···················· | ···················· | ···················· | ···················· | ···················· | |
| G | 6388 hCoV-19/USA/MN-MDH-1615/2020\|EPI_ISL_530211\|2020-0 | ···················· | ···················· | ···················· | ···················· | ···················· | ···················· | |
| G | 6389 hCoV-19/USA/MN-MDH-1616/2020\|EPI_ISL_530212\|2020-0 | ···················· | ···················· | ···················· | ···················· | ···················· | ···················· | |
| G | 6390 hCoV-19/USA/MN-MDH-1617/2020\|EPI_ISL_530213\|2020-0 | ···················· | ···················· | ···················· | ···················· | ···················· | ···················· | |
| G | 6391 hCoV-19/USA/MN-MDH-1619/2020\|EPI_ISL_530215\|2020-0 | ···················· | ···················· | ···················· | ···················· | ···················· | ···················· | |
| G | 6392 hCoV-19/USA/MN-MDH-1620/2020\|EPI_ISL_530216\|2020-0 | ···················· | ···················· | ···················· | ···················· | ···················· | ···················· | |
| G | 6393 hCoV-19/USA/MN-MDH-1621/2020\|EPI_ISL_530217\|2020-0 | ···················· | ···················· | ···················· | ···················· | ···················· | ···················· | |
| G | 6394 hCoV-19/USA/MN-MDH-1622/2020\|EPI_ISL_530218\|2020-0 | ···················· | ···················· | ···················· | ···················· | ···················· | ···················· | |
| G | 6395 hCoV-19/USA/MN-MDH-1623/2020\|EPI_ISL_530219\|2020-0 | ···················· | ···················· | ···················· | ···················· | ···················· | ···················· | |
| G | 6396 hCoV-19/USA/MN-MDH-1624/2020\|EPI_ISL_530220\|2020-0 | ···················· | ···················· | ···················· | ···················· | ···················· | ···················· | |
| G | 6397 hCoV-19/USA/MN-MDH-1625/2020\|EPI_ISL_530221\|2020-0 | ···················· | ···················· | ···················· | ···················· | ···················· | ···················· | |
| G | 6398 hCoV-19/USA/MN-MDH-1626/2020\|EPI_ISL_530222\|2020-0 | ···················· | ···················· | ···················· | ···················· | ···················· | ···················· | |
| G | 6399 hCoV-19/USA/MN-MDH-1627/2020\|EPI_ISL_530223\|2020-0 | ···················· | ···················· | ···················· | ···················· | ···················· | ···················· | |
| G | 6400 hCoV-19/USA/MN-MDH-1628/2020\|EPI_ISL_530224\|2020-0 | ···················· | ···················· | ···················· | ···················· | ···················· | ···················· | |
| G | 6405 hCoV-19/England/LIVE-A8732/2020\|EPI_ISL_517205\|202 | ···················· | ···················· | ···················· | ···················· | ·······NN··········· | ···················· | |
| G | 6409 hCoV-19/England/LIVE-A878D/2020\|EPI_ISL_517209\|202 | ···················· | ···················· | ···················· | ···················· | ···················· | ···················· | |
| G | 6410 hCoV-19/England/LIVE-A879C/2020\|EPI_ISL_517210\|202 | ···················· | ···················· | ···················· | ···················· | ···················· | ···················· | |
| G | 6411 hCoV-19/England/LIVE-A87AB/2020\|EPI_ISL_517211\|202 | ···················· | ···················· | ···················· | ···················· | ···················· | ···················· | |
| G | 6414 hCoV-19/England/LIVE-A87E7/2020\|EPI_ISL_517214\|202 | ···················· | ···················· | ···················· | ···················· | ···················· | ···················· | |
| G | 6415 hCoV-19/England/LIVE-A87F6/2020\|EPI_ISL_517215\|202 | ···················· | ···················· | ···················· | ···················· | ···················· | ···················· | |
| G | 6416 hCoV-19/England/LIVE-A8802/2020\|EPI_ISL_517216\|202 | ···················· | ···················· | ···················· | ···················· | ···················· | ···················· | |
| G | 6418 hCoV-19/England/LIVE-A8820/2020\|EPI_ISL_517218\|202 | ···················· | ···················· | ···················· | ···················· | ···················· | ···················· | |
| G | 6419 hCoV-19/England/LIVE-A883F/2020\|EPI_ISL_517219\|202 | ···················· | ···················· | ···················· | ···················· | ···················· | ···················· | |
| G | 6420 hCoV-19/England/LIVE-A884E/2020\|EPI_ISL_517220\|202 | ···················· | ···················· | ···················· | ···················· | ···················N | ···················· | 1 |
| G | 6421 hCoV-19/England/LIVE-A885D/2020\|EPI_ISL_517221\|202 | ···················· | ···················· | ···················· | ···················· | ···················· | ···················· | |
| G | 6422 hCoV-19/England/LIVE-A886C/2020\|EPI_ISL_517222\|202 | ···················· | ···················· | ···················· | ···················· | ···················· | ···················· | |
| G | 6423 hCoV-19/England/LIVE-A887B/2020\|EPI_ISL_517223\|202 | ···················· | ···················· | ···················· | ···················· | ···················· | ···················· | |
| G | 6424 hCoV-19/England/LIVE-A888A/2020\|EPI_ISL_517224\|202 | ···················· | ···················· | ···················· | ···················· | ···················· | ···················· | |
| G | 6426 hCoV-19/England/LIVE-A88A8/2020\|EPI_ISL_517226\|202 | ···················· | ···················· | ···················· | ···················· | ···················· | ···················· | |
| G | 6427 hCoV-19/England/LIVE-A88B7/2020\|EPI_ISL_517227\|202 | ···················· | ···················· | ···················· | ···················· | ···················· | ···················· | |
| G | 6429 hCoV-19/England/LIVE-A8F0D/2020\|EPI_ISL_517232\|202 | ···················· | ···················· | ···················· | ···················· | ···················· | ···················· | |

G 6431 hCoV-19/England/LIVE-A8F2B/2020|EPI_ISL_517234|202 ···················· ···················· ···················· ···················· ···················· ····················
G 6433 hCoV-19/England/LIVE-A8F58/2020|EPI_ISL_517236|202 ···················· ···················· ···················· ···················· ···················· ····················
G 6434 hCoV-19/England/LIVE-A8F67/2020|EPI_ISL_517237|202 ···················· ···················· ···················· ···················· ···················· ····················
G 6435 hCoV-19/England/LIVE-A8F76/2020|EPI_ISL_517238|202 ···················· ···················· ···················· ···················· ···················· ····················
G 6436 hCoV-19/England/LIVE-A8F94/2020|EPI_ISL_517240|202 ···················· ···················· ···················· ···················· ···················· ····················
G 6437 hCoV-19/England/LIVE-A8FA3/2020|EPI_ISL_517241|202 ···················· ···················· ···················· ···················· ···················· ····················
G 6438 hCoV-19/England/LIVE-A8FB2/2020|EPI_ISL_517242|202 ···················· ···················· ···················· ···················· ···················· ····················
G 6439 hCoV-19/England/LIVE-A8FC1/2020|EPI_ISL_517243|202 ···················· ···················· ···················· ···················· ···················· ····················
G 6441 hCoV-19/England/LIVE-A8FFE/2020|EPI_ISL_517246|202 ···················· ···················· ···················· ···················· ···················· ····················
G 6445 hCoV-19/England/LIVE-A9036/2020|EPI_ISL_517250|202 ···················· ·A·················· ···················· ···················· ···················· ····················
G 6446 hCoV-19/England/LIVE-A9045/2020|EPI_ISL_517251|202 ···················· ···················· ···················· ···················· ···················· ····················
G 6447 hCoV-19/England/LIVE-A9054/2020|EPI_ISL_517252|202 ···················· ···················· ···················· ···················· ···················· ····················
G 6448 hCoV-19/England/LIVE-A9063/2020|EPI_ISL_517253|202 ···················· ···················· ···················· ···················· ···················· ····················
G 6449 hCoV-19/England/LIVE-A9072/2020|EPI_ISL_517254|202 ···················· ···················· ···················· ···················· ···················· ····················
G 6450 hCoV-19/England/LIVE-A9081/2020|EPI_ISL_517255|202 ···T················ ···················· ···················· ···················· ···················· ····················
G 6451 hCoV-19/England/LIVE-A9090/2020|EPI_ISL_517256|202 ···················· ···················· ···················· ···················· ···················· ····················
G 6452 hCoV-19/England/LIVE-A90AF/2020|EPI_ISL_517257|202 ···················· ···················· ···················· ···················· ···················· ····················
G 6453 hCoV-19/England/LIVE-A90BE/2020|EPI_ISL_517258|202 ···················· ···················· ···················· ···················· ···················· ····················
G 6454 hCoV-19/England/LIVE-A90CD/2020|EPI_ISL_517259|202 ···················· ···················· ···················· ···················· ···················· ····················
G 6456 hCoV-19/England/LIVE-A90EB/2020|EPI_ISL_517261|202 ···················· ···················· ···················· ···················· ···················· ····················
G 6457 hCoV-19/England/LIVE-A9106/2020|EPI_ISL_517262|202 ···················· ···················· ···················· ···················· ···················· ····················
G 6458 hCoV-19/England/LIVE-A9115/2020|EPI_ISL_517263|202 ···················· ···················· ···················· ···················· ···················· ····················
G 6459 hCoV-19/England/LIVE-A9124/2020|EPI_ISL_517264|202 ···················· ···················· ···················· ···················· ···················· ····················
G 6461 hCoV-19/England/LIVE-A9142/2020|EPI_ISL_517266|202 ···················· ···················· ···················· ···················· ···················· ····················
G 6462 hCoV-19/England/LIVE-A9151/2020|EPI_ISL_517267|202 ···················· ···················· ···················· ···················· ···················· ····················
G 6463 hCoV-19/England/LIVE-A9160/2020|EPI_ISL_517268|202 ···················· ···················· ···················· ···················· ···················· ····················
G 6464 hCoV-19/England/LIVE-A917F/2020|EPI_ISL_517269|202 ···················· ···················· ···················· ···················· ···················· ····················
G 6465 hCoV-19/Australia/VIC5729/2020|EPI_ISL_518530|2020 ···················· ···················· ···················· ···················· ···················· ····················
G 6466 hCoV-19/Australia/VIC5730/2020|EPI_ISL_518531|2020 ···················· ···················· ···················· ···················· ···················· ····················
G 6467 hCoV-19/Australia/VIC5732/2020|EPI_ISL_518533|2020 ···················· ···················· ···················· ···················· ···················· ····················
G 6468 hCoV-19/Australia/VIC5733/2020|EPI_ISL_518534|2020 ···················· ···················· ···················· ···················· ···················· ····················
G 6469 hCoV-19/Australia/VIC5734/2020|EPI_ISL_518535|2020 ···················· ···················· ···················· ···················· ···················· ····················
G 6470 hCoV-19/Australia/VIC5735/2020|EPI_ISL_518536|2020 ···················· ···················· ···················· ···················· ···················· ····················
G 6471 hCoV-19/Australia/VIC5736/2020|EPI_ISL_518537|2020 ···················· ···················· ···················· ···················· ···················· ····················
G 6472 hCoV-19/Australia/VIC5737/2020|EPI_ISL_518538|2020 ···················· ···················· ···················· ···················· ···················· ····················
G 6473 hCoV-19/Australia/VIC5738/2020|EPI_ISL_518539|2020 ···················· ···················· ···················· ···················· ···················· ····················
G 6474 hCoV-19/Australia/VIC5739/2020|EPI_ISL_518540|2020 ···················· ···················· ···················· ···················· ···················· ····················
G 6475 hCoV-19/Australia/VIC5740/2020|EPI_ISL_518541|2020 ···················· ···················· ···················· ···················· ···················· ····················
G 6476 hCoV-19/Australia/VIC5741/2020|EPI_ISL_518542|2020 ···················· ···················· ···················· ···················· ···················· ····················
G 6477 hCoV-19/Australia/VIC5743/2020|EPI_ISL_518544|2020 ···················· ···················· ···················· ···················· ···················· ····················
G 6478 hCoV-19/Australia/VIC5746/2020|EPI_ISL_518545|2020 ···················· ···················· ···················· ···················· ···················· ····················
G 6479 hCoV-19/Australia/VIC5747/2020|EPI_ISL_518546|2020 ···················· ···················· ···················· ···················· ···················· ····················
G 6480 hCoV-19/Australia/VIC5748/2020|EPI_ISL_518547|2020 ···················· ···················· ···················· ···················· ···················· ····················
G 6481 hCoV-19/Australia/VIC5749/2020|EPI_ISL_518548|2020 ···················· ···················· ···················· ···················· ···················· ····················
G 6482 hCoV-19/Australia/VIC5750/2020|EPI_ISL_518549|2020 ···················· ···················· ···················· ···················· ···················· ····················
G 6483 hCoV-19/Australia/VIC5752/2020|EPI_ISL_518550|2020 ···················· ···················· ···················· ···················· ···················· ····················
G 6484 hCoV-19/Australia/VIC5753/2020|EPI_ISL_518551|2020 ···················· ···················· ···················· ···················· ···················· ····················
G 6485 hCoV-19/Australia/VIC5754/2020|EPI_ISL_518552|2020 ···················· ···················· ···················· ···················· ···················· ····················
G 6486 hCoV-19/Australia/VIC5755/2020|EPI_ISL_518553|2020 ···················· ···················· ···················· ···················· ···················· ····················
G 6487 hCoV-19/Australia/VIC5756/2020|EPI_ISL_518554|2020 ···················· ···················· ···················· ···················· ···················· ····················
G 6488 hCoV-19/Australia/VIC5757/2020|EPI_ISL_518555|2020 ···················· ···················· ···················· ···················· ···················· ····················
G 6489 hCoV-19/Australia/VIC5758/2020|EPI_ISL_518556|2020 ···················· ···················· ···················· ···················· ···················· ····················
G 6490 hCoV-19/Australia/VIC5759/2020|EPI_ISL_518557|2020 ···················· ···················· ···················· ···················· ···················· ····················
G 6491 hCoV-19/Australia/VIC5760/2020|EPI_ISL_518558|2020 ···················· ···················· ···················· ···················· ···················· ····················
G 6492 hCoV-19/Australia/VIC5761/2020|EPI_ISL_518559|2020 ···················· ···················· ···················· ···················· ···················· ····················
G 6493 hCoV-19/Australia/VIC5762/2020|EPI_ISL_518560|2020 ···················· ···················· ···················· ···················· ···················· ····················
G 6494 hCoV-19/Australia/VIC5763/2020|EPI_ISL_518561|2020 ···················· ···················· ···················· ···················· ···················· ····················
G 6495 hCoV-19/Australia/VIC5764/2020|EPI_ISL_518562|2020 ···················· ···················· ···················· ···················· ···················· ····················
G 6496 hCoV-19/Australia/VIC5765/2020|EPI_ISL_518563|2020 ···················· ···················· ···················· ···················· ···················· ····················
G 6497 hCoV-19/Australia/VIC5766/2020|EPI_ISL_518564|2020 ···················· ···················· ···················· ···················· ···················· ····················
G 6498 hCoV-19/Australia/VIC5767/2020|EPI_ISL_518565|2020 ···················· ···················· ···················· ···················· ···················· ····················
G 6499 hCoV-19/Australia/VIC5769/2020|EPI_ISL_518567|2020 ···················· ···················· ···················· ···················· ···················· ····················
G 6500 hCoV-19/Australia/VIC5770/2020|EPI_ISL_518568|2020 ···················· ···················· ···················· ···················· ···················· ····················
G 6501 hCoV-19/Australia/VIC5771/2020|EPI_ISL_518569|2020 ···················· ···················· ···················· ···················· ···················· ····················
G 6502 hCoV-19/Australia/VIC5772/2020|EPI_ISL_518570|2020 ···················· ···················· ···················· ···················· ···················· ····················
G 6503 hCoV-19/Australia/VIC5773/2020|EPI_ISL_518571|2020 ···················· ···················· ···················· ···················· ···················· ····················
G 6504 hCoV-19/Australia/VIC5774/2020|EPI_ISL_518572|2020 ···················· ···················· ···················· ···················· ···················· ····················
G 6505 hCoV-19/Australia/VIC5775/2020|EPI_ISL_518573|2020 ···················· ···················· ···················· ···················· ···················· ····················
G 6506 hCoV-19/Australia/VIC5776/2020|EPI_ISL_518574|2020 ···················· ···················· ···················· ···················· ···················· ····················
G 6507 hCoV-19/Australia/VIC5778/2020|EPI_ISL_518576|2020 ···················· ···················· ···················· ···················· ···················· ····················
G 6508 hCoV-19/Australia/VIC5779/2020|EPI_ISL_518577|2020 ···················· ···················· ···················· ···················· ···················· ····················
G 6509 hCoV-19/Australia/VIC5780/2020|EPI_ISL_518578|2020 ···················· ···················· ···················· ···················· ···················· ····················
G 6510 hCoV-19/Australia/VIC5781/2020|EPI_ISL_518579|2020 ···················· ···················· ···················· ···················· ···················· ····················
G 6511 hCoV-19/Australia/VIC5782/2020|EPI_ISL_518580|2020 ···················· ···················· ···················· ···················· ···················· ····················
G 6512 hCoV-19/Australia/VIC5784/2020|EPI_ISL_518581|2020 ···················· ···················· ···················· ···················· ···················· ····················
G 6513 hCoV-19/Australia/VIC5785/2020|EPI_ISL_518582|2020 ···················· ···················· ···················· ···················· ···················· ····················
G 6514 hCoV-19/Australia/VIC5787/2020|EPI_ISL_518584|2020 ···················· ···················· ···················· ···················· ···················· ····················
G 6515 hCoV-19/Australia/VIC5790/2020|EPI_ISL_518586|2020 ···················· ···················· ···················· ···················· ···················· ····················
G 6516 hCoV-19/Australia/VIC5791/2020|EPI_ISL_518587|2020 ···················· ···················· ···················· ···················· ···················· ····················
G 6517 hCoV-19/Australia/VIC5792/2020|EPI_ISL_518588|2020 ···················· ···················· ···················· ···················· ···················· ····················

| | | | | | | | |
|---|---|---|---|---|---|---|---|
| G | 6518 hCoV-19/Australia/VIC5793/2020\|EPI_ISL_518589\|2020 | ···················· | ···················· | ···················· | ···················· | ···················· | ···················· |
| G | 6519 hCoV-19/Australia/VIC5794/2020\|EPI_ISL_518590\|2020 | ···················· | ···················· | ···················· | ···················· | ···················· | ···················· |
| G | 6520 hCoV-19/Australia/VIC5795/2020\|EPI_ISL_518591\|2020 | ···················· | ···················· | ···················· | ···················· | ···················· | ···················· |
| G | 6521 hCoV-19/Australia/VIC5796/2020\|EPI_ISL_518592\|2020 | ···················· | ···················· | ···················· | ···················· | ···················· | ···················· |
| G | 6522 hCoV-19/Australia/VIC5799/2020\|EPI_ISL_518594\|2020 | ···················· | ···················· | ···················· | ···················· | ···················· | ···················· |
| G | 6523 hCoV-19/Australia/VIC5800/2020\|EPI_ISL_518595\|2020 | ···················· | ···················· | ···················· | ···················· | ···················· | ···················· |
| G | 6524 hCoV-19/Australia/VIC5801/2020\|EPI_ISL_518596\|2020 | ···················· | ···················· | ···················· | ···················· | ···················· | ···················· |
| G | 6525 hCoV-19/Australia/VIC5802/2020\|EPI_ISL_518597\|2020 | ···················· | ···················· | ···················· | ···················· | ···················· | ···················· |
| G | 6526 hCoV-19/Australia/VIC5804/2020\|EPI_ISL_518599\|2020 | ···················· | ···················· | ···················· | ···················· | ···················· | ···················· |
| G | 6527 hCoV-19/Australia/VIC5057/2020\|EPI_ISL_519860\|2020 | ···················· | ···················· | ···················· | ···················· | ···················· | ···················· |
| G | 6528 hCoV-19/Australia/VIC5058/2020\|EPI_ISL_519861\|2020 | ···················· | ···················· | ···················· | ···················· | ···················· | ···················· |
| G | 6529 hCoV-19/Australia/VIC5059/2020\|EPI_ISL_519862\|2020 | ···················· | ···················· | ···················· | ···················· | ···················· | ···················· |
| G | 6530 hCoV-19/Australia/VIC5060/2020\|EPI_ISL_519863\|2020 | ···················· | ···················· | ···················· | ···················· | ···················· | ···················· |
| G | 6531 hCoV-19/Australia/VIC5061/2020\|EPI_ISL_519864\|2020 | ···················· | ···················· | ···················· | ···················· | ···················· | ···················· |
| G | 6532 hCoV-19/Australia/VIC5062/2020\|EPI_ISL_519865\|2020 | ···················· | ···················· | ···················· | ···················· | ···················· | ···················· |
| G | 6533 hCoV-19/Australia/VIC5063/2020\|EPI_ISL_519866\|2020 | ···················· | ···················· | ···················· | ···················· | ···················· | ···················· |
| G | 6534 hCoV-19/Australia/VIC5064/2020\|EPI_ISL_519867\|2020 | ···················· | ···················· | ···················· | ···················· | ···················· | ···················· |
| G | 6535 hCoV-19/Australia/VIC5065/2020\|EPI_ISL_519868\|2020 | ···················· | ···················· | ···················· | ···················· | ···················· | ···················· |
| G | 6536 hCoV-19/Australia/VIC5066/2020\|EPI_ISL_519869\|2020 | ···················· | ···················· | ···················· | ···················· | ···················· | ···················· |
| G | 6537 hCoV-19/Australia/VIC5067/2020\|EPI_ISL_519870\|2020 | ···················· | ···················· | ···················· | ···················· | ···················· | ···················· |
| G | 6538 hCoV-19/Australia/VIC5068/2020\|EPI_ISL_519871\|2020 | ···················· | ···················· | ···················· | ···················· | ···················· | ···················· |
| G | 6539 hCoV-19/Australia/VIC5069/2020\|EPI_ISL_519872\|2020 | ···················· | ···················· | ···················· | ···················· | ···················· | ···················· |
| G | 6540 hCoV-19/Australia/VIC5070/2020\|EPI_ISL_519873\|2020 | ···················· | ···················· | ···················· | ···················· | ···················· | ···················· |
| G | 6541 hCoV-19/Australia/VIC5071/2020\|EPI_ISL_519874\|2020 | ···················· | ···················· | ···················· | ···················· | ···················· | ···················· |
| G | 6542 hCoV-19/Australia/VIC5072/2020\|EPI_ISL_519875\|2020 | ···················· | ···················· | ···················· | ···················· | ···················· | ···················· |
| G | 6543 hCoV-19/Australia/VIC5073/2020\|EPI_ISL_519876\|2020 | ···················· | ···················· | ···················· | ···················· | ···················· | ···················· |
| G | 6544 hCoV-19/Australia/VIC5074/2020\|EPI_ISL_519877\|2020 | ···················· | ···················· | ···················· | ···················· | ···················· | ···················· |
| G | 6545 hCoV-19/Australia/VIC5075/2020\|EPI_ISL_519878\|2020 | ···················· | ···················· | ···················· | ···················· | ···················· | ···················· |
| G | 6546 hCoV-19/Australia/VIC5077/2020\|EPI_ISL_519880\|2020 | ···················· | ···················· | ···················· | ···················· | ···················· | ···················· |
| G | 6547 hCoV-19/Australia/VIC5078/2020\|EPI_ISL_519881\|2020 | ···················· | ···················· | ···················· | ···················· | ···················· | ···················· |
| G | 6548 hCoV-19/Australia/VIC5079/2020\|EPI_ISL_519882\|2020 | ···················· | ···················· | ···················· | ···················· | ···················· | ···················· |
| G | 6549 hCoV-19/Australia/VIC5080/2020\|EPI_ISL_519883\|2020 | ···················· | ···················· | ···················· | ···················· | ···················· | ···················· |
| G | 6550 hCoV-19/Australia/VIC5081/2020\|EPI_ISL_519884\|2020 | ···················· | ···················· | ···················· | ···················· | ···················· | ···················· |
| G | 6551 hCoV-19/Australia/VIC5082/2020\|EPI_ISL_519885\|2020 | ···················· | ···················· | ···················· | ···················· | ···················· | ···················· |
| G | 6552 hCoV-19/Australia/VIC5083/2020\|EPI_ISL_519886\|2020 | ···················· | ···················· | ···················· | ···················· | ···················· | ···················· |
| G | 6553 hCoV-19/Australia/VIC5084/2020\|EPI_ISL_519887\|2020 | ···················· | ···················· | ···················· | ···················· | ···················· | ···················· |
| G | 6554 hCoV-19/Australia/VIC5085/2020\|EPI_ISL_519888\|2020 | ···················· | ···················· | ···················· | ···················· | ···················· | ···················· |
| G | 6555 hCoV-19/Australia/VIC5086/2020\|EPI_ISL_519889\|2020 | ···················· | ···················· | ···················· | ···················· | ···················· | ···················· |
| G | 6556 hCoV-19/Australia/VIC5087/2020\|EPI_ISL_519890\|2020 | ···················· | ···················· | ···················· | ···················· | ···················· | ···················· |
| G | 6557 hCoV-19/Australia/VIC5088/2020\|EPI_ISL_519891\|2020 | ···················· | ···················· | ···················· | ···················· | ···················· | ···················· |
| G | 6558 hCoV-19/Australia/VIC5089/2020\|EPI_ISL_519892\|2020 | ···················· | ···················· | ···················· | ···················· | ···················· | ···················· |
| G | 6559 hCoV-19/Australia/VIC5090/2020\|EPI_ISL_519893\|2020 | ···················· | ···················· | ···················· | ···················· | ···················· | ···················· |
| G | 6560 hCoV-19/Australia/VIC5091/2020\|EPI_ISL_519894\|2020 | ···················· | ···················· | ···················· | ···················· | ···················· | ···················· |
| G | 6561 hCoV-19/Australia/VIC5093/2020\|EPI_ISL_519896\|2020 | ···················· | ···················· | ···················· | ···················· | ···················· | ···················· |
| G | 6562 hCoV-19/Australia/VIC5094/2020\|EPI_ISL_519897\|2020 | ···················· | ···················· | ···················· | ···················· | ···················· | ···················· |
| G | 6563 hCoV-19/Australia/VIC5095/2020\|EPI_ISL_519898\|2020 | ···················· | ···················· | ···················· | ···················· | ···················· | ···················· |
| G | 6564 hCoV-19/Australia/VIC5096/2020\|EPI_ISL_519899\|2020 | ···················· | ···················· | ···················· | ···················· | ···················· | ···················· |
| G | 6565 hCoV-19/Australia/VIC3134/2020\|EPI_ISL_520841\|2020 | ···················· | ···················· | ···················· | ···················· | ··N················· | ···················· |
| G | 6566 hCoV-19/Australia/VIC3135/2020\|EPI_ISL_520842\|2020 | ···················· | ···················· | ···················· | ···················· | ···················· | ···················· |
| G | 6567 hCoV-19/Australia/VIC3136/2020\|EPI_ISL_520843\|2020 | ···················· | ···················· | ···················· | ···················· | ···················· | ···················· |
| G | 6568 hCoV-19/Australia/VIC3137/2020\|EPI_ISL_520844\|2020 | ···················· | ···················· | ···················· | ···················· | ···················· | ···················· |
| G | 6570 hCoV-19/Australia/VIC3139/2020\|EPI_ISL_520846\|2020 | ···················· | ···················· | ···················· | ···················· | ···················· | ···················· |
| G | 6571 hCoV-19/Australia/VIC3140/2020\|EPI_ISL_520847\|2020 | ···················· | ···················· | ···················· | ···················· | ···················· | ···················· |
| G | 6572 hCoV-19/Australia/VIC3141/2020\|EPI_ISL_520848\|2020 | ···················· | ···················· | ···················· | ···················· | ···················· | ···················· |
| G | 6573 hCoV-19/Australia/VIC3142/2020\|EPI_ISL_520849\|2020 | ···················· | ···················· | ···················· | ···················· | ···················· | ···················· |
| G | 6574 hCoV-19/Australia/VIC3143/2020\|EPI_ISL_520850\|2020 | ···················· | ···················· | ···················· | ···················· | ···················· | ···················· |
| G | 6575 hCoV-19/Australia/VIC3144/2020\|EPI_ISL_520851\|2020 | ···················· | ···················· | ···················· | ···················· | ···················· | ···················· |
| G | 6577 hCoV-19/Australia/VIC3146/2020\|EPI_ISL_520853\|2020 | ···················· | ···················· | ···················· | ···················· | ···················· | ···················· |
| G | 6579 hCoV-19/Australia/VIC3149/2020\|EPI_ISL_520855\|2020 | ···················· | ···················· | ···················· | ···················· | ···················· | ···················· |
| G | 6580 hCoV-19/Australia/VIC3150/2020\|EPI_ISL_520856\|2020 | ···················· | ···················· | ···················· | ···················· | ···················· | ···················· |
| G | 6582 hCoV-19/Australia/VIC3152/2020\|EPI_ISL_520858\|2020 | ···················· | ···················· | ···················· | ···················· | ·NN················· | ···················· |
| G | 6583 hCoV-19/Australia/VIC3153/2020\|EPI_ISL_520859\|2020 | ···················· | ···················· | ···················· | ···················· | ···················· | ···················· |
| G | 6584 hCoV-19/Australia/VIC3154/2020\|EPI_ISL_520860\|2020 | ···················· | ···················· | ···················· | ···················· | ···················· | ···················· |
| G | 6585 hCoV-19/Australia/VIC3155/2020\|EPI_ISL_520861\|2020 | ···················· | ···················· | ···················· | ···················· | ···················· | ···················· |
| G | 6586 hCoV-19/Australia/VIC3156/2020\|EPI_ISL_520862\|2020 | ···················· | ···················· | ···················· | ···················· | ···················· | ···················· |
| G | 6587 hCoV-19/Australia/VIC3157/2020\|EPI_ISL_520863\|2020 | ···················· | ···················· | ···················· | ···················· | ···················· | ···················· |
| G | 6588 hCoV-19/Australia/VIC3172/2020\|EPI_ISL_520864\|2020 | ···················· | ···················· | ···················· | ···················· | ···················· | ···················· |
| G | 6589 hCoV-19/Australia/VIC3173/2020\|EPI_ISL_520865\|2020 | ···················· | ···················· | ···················· | ···················· | ···················· | ···················· |
| G | 6590 hCoV-19/Australia/VIC3174/2020\|EPI_ISL_520866\|2020 | ···················· | ···················· | ···················· | ···················· | ···················· | ···················· |
| G | 6591 hCoV-19/Australia/VIC3176/2020\|EPI_ISL_520867\|2020 | ···················· | ···················· | ···················· | ···················· | ···················· | ···················· |
| G | 6592 hCoV-19/Australia/VIC3177/2020\|EPI_ISL_520868\|2020 | ···················· | ···················· | ···················· | ···················· | ···················· | ···················· |
| G | 6593 hCoV-19/Australia/VIC3178/2020\|EPI_ISL_520869\|2020 | ···················· | ···················· | ···················· | ···················· | ···················· | ···················· |
| G | 6594 hCoV-19/Australia/VIC3179/2020\|EPI_ISL_520870\|2020 | ···················· | ···················· | ···················· | ···················· | ···················· | ···················· |
| G | 6595 hCoV-19/Australia/VIC3181/2020\|EPI_ISL_520871\|2020 | ···················· | ···················· | ···················· | ···················· | ···················· | ···················· |
| G | 6596 hCoV-19/Australia/VIC3182/2020\|EPI_ISL_520872\|2020 | ···················· | ···················· | ···················· | ···················· | ···················· | ···················· |
| G | 6597 hCoV-19/Australia/VIC3186/2020\|EPI_ISL_520873\|2020 | ···················· | ···················· | ···················· | ···················· | ···················· | ···················· |
| G | 6598 hCoV-19/Australia/VIC3187/2020\|EPI_ISL_520874\|2020 | ···················· | ···················· | ···················· | ···················· | ···················· | ···················· |
| G | 6599 hCoV-19/Australia/VIC3188/2020\|EPI_ISL_520875\|2020 | ···················· | ···················· | ···················· | ···················· | ···················· | ···················· |
| G | 6600 hCoV-19/Australia/VIC3189/2020\|EPI_ISL_520876\|2020 | ···················· | ···················· | ···················· | ···················· | ···················· | ···················· |
| G | 6601 hCoV-19/Australia/VIC3190/2020\|EPI_ISL_520877\|2020 | ···················· | ···················· | ···················· | ···················· | ···················· | ···················· |

| | | | | | | | | | |
|---|---|---|---|---|---|---|---|---|---|
| G | 6602 | hCoV-19/Australia/VIC3192/2020\|EPI_ISL_520878\|2020 | ···················· | ···················· | ···················· | ···················· | ···················· | ···················· | |
| G | 6603 | hCoV-19/Australia/VIC3193/2020\|EPI_ISL_520879\|2020 | ···················· | ···················· | ···················· | ···················· | ···················· | ···················· | |
| G | 6604 | hCoV-19/Australia/VIC3194/2020\|EPI_ISL_520880\|2020 | ···················· | ···················· | ···················· | ···················· | ···················· | ···················· | |
| G | 6605 | hCoV-19/Australia/VIC3195/2020\|EPI_ISL_520881\|2020 | ···················· | ···················· | ···················· | ···················· | ···················· | ···················· | |
| G | 6606 | hCoV-19/Australia/VIC3196/2020\|EPI_ISL_520882\|2020 | ···················· | ···················· | ···················· | ···················· | ···················· | ···················· | |
| G | 6607 | hCoV-19/Australia/VIC3197/2020\|EPI_ISL_520883\|2020 | ···················· | ···················· | ···················· | ···················· | NNN················· | ···················· | 1 |
| G | 6608 | hCoV-19/Australia/VIC3198/2020\|EPI_ISL_520884\|2020 | ···················· | ···················· | ···················· | ···················· | ···················· | ···················· | |
| G | 6609 | hCoV-19/Australia/VIC3199/2020\|EPI_ISL_520885\|2020 | ···················· | ···················· | ···················· | ···················· | ···················· | ···················· | |
| G | 6610 | hCoV-19/Australia/VIC3200/2020\|EPI_ISL_520886\|2020 | ···················· | ···················· | ···················· | ···················· | ···················· | ···················· | |
| G | 6611 | hCoV-19/Australia/VIC3201/2020\|EPI_ISL_520887\|2020 | ···················· | ···················· | ···················· | ···················· | ···················· | ···················· | |
| G | 6612 | hCoV-19/Australia/VIC3202/2020\|EPI_ISL_520888\|2020 | ···················· | ···················· | ···················· | ···················· | ···················· | ···················· | |
| G | 6613 | hCoV-19/Australia/VIC3203/2020\|EPI_ISL_520889\|2020 | ···················· | ···················· | ···················· | ···················· | ···················· | ···················· | |
| G | 6614 | hCoV-19/Australia/VIC3204/2020\|EPI_ISL_520890\|2020 | ···················· | ···················· | ···················· | ···················· | ···················· | ···················· | |
| G | 6615 | hCoV-19/Australia/VIC3205/2020\|EPI_ISL_520891\|2020 | ···················· | ···················· | ···················· | ···················· | ···················· | ···················· | |
| G | 6616 | hCoV-19/Australia/VIC3206/2020\|EPI_ISL_520892\|2020 | ···················· | ···················· | ···················· | ···················· | ···················· | ···················· | |
| G | 6617 | hCoV-19/Australia/VIC3207/2020\|EPI_ISL_520893\|2020 | ···················· | ···················· | ···················· | ···················· | ···················· | ···················· | |
| G | 6619 | hCoV-19/Australia/VIC3209/2020\|EPI_ISL_520895\|2020 | ···················· | ···················· | ···················· | ···················· | ···················· | ···················· | |
| G | 6620 | hCoV-19/Australia/VIC3210/2020\|EPI_ISL_520896\|2020 | ···················· | ···················· | ···················· | ···················· | ···················· | ···················· | |
| G | 6621 | hCoV-19/Australia/VIC3211/2020\|EPI_ISL_520897\|2020 | ···················· | ···················· | ···················· | ···················· | ···················· | ···················· | |
| G | 6622 | hCoV-19/Australia/VIC3212/2020\|EPI_ISL_520898\|2020 | ···················· | ···················· | ···················· | ···················· | ···················· | ···················· | |
| G | 6623 | hCoV-19/Australia/VIC3214/2020\|EPI_ISL_520899\|2020 | ···················· | ···················· | ···················· | ···················· | ···················· | ···················· | |
| G | 6624 | hCoV-19/USA/VA-DCLS-0605/2020\|EPI_ISL_526822\|2020- | ···················· | ···················· | ···················· | ···················· | ···················· | ···················· | |
| G | 6625 | hCoV-19/USA/SC-DHEC-0588/2020\|EPI_ISL_529097\|2020- | ···················· | ···················· | ···················· | ···················· | ···················· | ···················· | |
| G | 6626 | hCoV-19/USA/SC-DHEC-0589/2020\|EPI_ISL_529098\|2020- | ···················· | ···················· | ···················· | ···················· | ···················· | ···················· | |
| G | 6627 | hCoV-19/USA/SC-DHEC-0590/2020\|EPI_ISL_529099\|2020- | ···················· | ···················· | ···················· | ···················· | ···················· | ···················· | |
| G | 6628 | hCoV-19/England/BIRM-61AC3/2020\|EPI_ISL_529218\|202 | ···················· | ···················· | ···················· | ···················· | ···················· | ···················· | |
| G | 6629 | hCoV-19/England/BIRM-61B57/2020\|EPI_ISL_529219\|202 | ···················· | ···················· | ···················· | ···················· | ···················· | ···················· | |
| G | 6630 | hCoV-19/England/BIRM-61BB1/2020\|EPI_ISL_529220\|202 | ···················· | ···················· | ···················· | ···················· | ···················· | ···················· | |
| G | 6631 | hCoV-19/England/BIRM-61C09/2020\|EPI_ISL_529221\|202 | ···················· | ···················· | ···················· | ···················· | ···················· | ···················· | |
| G | 6632 | hCoV-19/England/BIRM-61E4F/2020\|EPI_ISL_529222\|202 | ···················· | ···················· | ···················· | ···················· | ···················· | ···················· | |
| G | 6633 | hCoV-19/England/BIRM-61EE5/2020\|EPI_ISL_529223\|202 | ···················· | ···················· | ···················· | ···················· | ···················· | ···················· | |
| G | 6634 | hCoV-19/England/CAMB-1B5BDB/2020\|EPI_ISL_529224\|20 | ···················· | ···················· | ···················· | ···················· | ···················· | ···················· | |
| G | 6635 | hCoV-19/England/CAMB-1B5CAB/2020\|EPI_ISL_529225\|20 | ···················· | ···················· | ···················· | ···················· | ···················· | ···················· | |
| G | 6636 | hCoV-19/Wales/PHWC-169C97/2020\|EPI_ISL_529226\|2020 | ···················· | ···················· | ···················· | ···················· | ···················· | ···················· | |
| G | 6637 | hCoV-19/Wales/PHWC-169E91/2020\|EPI_ISL_529227\|2020 | ···················· | ···················· | ···················· | ···················· | ···················· | ···················· | |
| G | 6638 | hCoV-19/Wales/PHWC-169ECE/2020\|EPI_ISL_529228\|2020 | ···················· | ···················· | ···················· | ···················· | ···················· | ···················· | |
| G | 6639 | hCoV-19/Wales/PHWC-169EEC/2020\|EPI_ISL_529229\|2020 | ···················· | ···················· | ···················· | ···················· | ···················· | ···················· | |
| G | 6640 | hCoV-19/Wales/PHWC-169EFB/2020\|EPI_ISL_529230\|2020 | ···················· | ···················· | ···················· | ···················· | ···················· | ···················· | |
| G | 6641 | hCoV-19/Wales/PHWC-169F16/2020\|EPI_ISL_529231\|2020 | ···················· | ···················· | ···················· | ···················· | ···················· | ···················· | |
| G | 6642 | hCoV-19/Wales/PHWC-169F43/2020\|EPI_ISL_529232\|2020 | ···················· | ···················· | ···················· | ···················· | ···················· | ···················· | |
| G | 6643 | hCoV-19/Wales/PHWC-169FCB/2020\|EPI_ISL_529233\|2020 | ···················· | ···················· | ···················· | ···················· | ···················· | ···················· | |
| G | 6644 | hCoV-19/England/BIRM-602BD/2020\|EPI_ISL_529234\|202 | ···················· | ···················· | ···················· | ···················· | ···················· | ···················· | |
| G | 6645 | hCoV-19/England/BIRM-635DF/2020\|EPI_ISL_529235\|202 | ···················· | ···················· | ···················· | ···················· | ···················· | ···················· | |
| G | 6646 | hCoV-19/England/CAMB-1B5BCC/2020\|EPI_ISL_529236\|20 | ···················· | ···················· | ···················· | ···················· | ···················· | ···················· | |
| G | 6647 | hCoV-19/England/BIRM-623D6/2020\|EPI_ISL_529237\|202 | ···················· | ···················· | ···················· | ···················· | ···················· | ···················· | |
| G | 6648 | hCoV-19/England/NORW-EEACE/2020\|EPI_ISL_529238\|202 | ···················· | ···················· | ···················· | ···················· | ···················· | ···················· | |
| G | 6649 | hCoV-19/England/BIRM-61841/2020\|EPI_ISL_529239\|202 | ···················· | ···················· | ···················· | ···················· | ···················· | ···················· | |
| G | 6650 | hCoV-19/England/NORW-EE563/2020\|EPI_ISL_529240\|202 | ···················· | ···················· | ···················· | ···················· | ···················· | ···················· | |
| G | 6651 | hCoV-19/England/NORW-EE581/2020\|EPI_ISL_529241\|202 | ···················· | ···················· | ···················· | ···················· | ···················· | ···················· | |
| G | 6652 | hCoV-19/England/NORW-EE5AF/2020\|EPI_ISL_529242\|202 | ···················· | ···················· | ···················· | ···················· | ···················· | ···················· | |
| G | 6653 | hCoV-19/England/NORW-EE730/2020\|EPI_ISL_529243\|202 | ···················· | ···················· | ···················· | ···················· | ···················· | ···················· | |
| G | 6654 | hCoV-19/England/NORW-EE77C/2020\|EPI_ISL_529245\|202 | ···················· | ···················· | ···················· | ···················· | ···················· | ···················· | |
| G | 6655 | hCoV-19/England/NORW-EEAA0/2020\|EPI_ISL_529248\|202 | ···················· | ···················· | ···················· | ···················· | ···················· | ···················· | |
| G | 6656 | hCoV-19/England/NORW-EEB16/2020\|EPI_ISL_529249\|202 | ···················· | ···················· | ···················· | ···················· | ···················· | ···················· | |
| G | 6661 | hCoV-19/England/LIVE-A7609/2020\|EPI_ISL_517047\|202 | ···················· | ···················· | ···················· | ···················· | ···················· | ···················· | |
| G | 6662 | hCoV-19/England/LIVE-A7618/2020\|EPI_ISL_517048\|202 | ···················· | ···················· | ···················· | ···················· | ···················· | N··················· | |
| G | 6667 | hCoV-19/England/LIVE-A76AF/2020\|EPI_ISL_517055\|202 | ···················· | ···················· | ···················· | ···················· | ···················· | ···················· | |
| G | 6668 | hCoV-19/England/LIVE-A76CD/2020\|EPI_ISL_517057\|202 | ···················· | ···················· | ···················· | ···················· | ···················· | ···················· | |
| G | 6669 | hCoV-19/England/LIVE-A76DC/2020\|EPI_ISL_517058\|202 | ···················· | ···················· | ···················· | ···················· | ···················· | ···················· | |
| G | 6670 | hCoV-19/England/LIVE-A76EB/2020\|EPI_ISL_517059\|202 | ···················· | ···················· | ···················· | ···················· | ···················· | ···················· | |
| G | 6672 | hCoV-19/England/LIVE-A7706/2020\|EPI_ISL_517061\|202 | ···················· | ···················· | ···················· | ···················· | ···················· | ···················· | |
| G | 6673 | hCoV-19/England/LIVE-A7733/2020\|EPI_ISL_517063\|202 | ···················· | ···················· | ···················· | ···················· | ···················· | ···················· | |
| G | 6674 | hCoV-19/England/LIVE-A7742/2020\|EPI_ISL_517064\|202 | ···················· | ···················· | ···················· | ···················· | ···················· | ···················· | |
| G | 6676 | hCoV-19/England/LIVE-A778E/2020\|EPI_ISL_517068\|202 | ···················· | ···················· | ···················· | ···················· | ···················· | ···················· | |
| G | 6677 | hCoV-19/England/LIVE-A77BB/2020\|EPI_ISL_517070\|202 | ···················· | ···················· | ···················· | ···················· | ······NN············ | ···················· | |
| G | 6678 | hCoV-19/England/LIVE-A77CA/2020\|EPI_ISL_517071\|202 | ···················· | ···················· | ···················· | ···················· | ···················· | ···················· | |
| G | 6681 | hCoV-19/England/LIVE-A77F7/2020\|EPI_ISL_517074\|202 | ···················· | ···················· | ···················· | ···················· | ···················· | ···················· | |
| G | 6682 | hCoV-19/England/LIVE-A7803/2020\|EPI_ISL_517075\|202 | ···················· | ···················· | ···················· | ···················· | ···················· | ···················· | |
| G | 6683 | hCoV-19/England/LIVE-A7812/2020\|EPI_ISL_517076\|202 | ···················· | ···················· | ···················· | ···················· | ···················· | ···················· | |
| G | 6686 | hCoV-19/England/LIVE-A784F/2020\|EPI_ISL_517079\|202 | ···················· | ···················· | ···················· | ···················· | ···················· | ···················· | |
| G | 6687 | hCoV-19/England/LIVE-A786D/2020\|EPI_ISL_517080\|202 | ···················· | ···················· | ···················· | ···················· | ···················· | ···················· | |
| G | 6688 | hCoV-19/England/LIVE-A787C/2020\|EPI_ISL_517081\|202 | ···················· | ·A·················· | ···················· | ···················· | ···················· | ···················· | |
| G | 6689 | hCoV-19/England/LIVE-A788B/2020\|EPI_ISL_517082\|202 | ···················· | ···················· | ···················· | ···················· | ···················· | ···················· | |
| G | 6690 | hCoV-19/England/LIVE-A789A/2020\|EPI_ISL_517083\|202 | ···················· | ···················· | ···················· | ···················· | ···················· | ···················· | |
| G | 6693 | hCoV-19/England/LIVE-A78D6/2020\|EPI_ISL_517087\|202 | ···················· | ···················· | ···················· | ···················· | ···················· | ···················· | |
| G | 6696 | hCoV-19/Australia/VIC5539/2020\|EPI_ISL_518370\|2020 | ···················· | ···················· | ···················· | ···················· | ···················· | ···················· | |
| G | 6697 | hCoV-19/Australia/VIC5540/2020\|EPI_ISL_518371\|2020 | ···················· | ···················· | ···················· | ···················· | ···················· | ···················· | |
| G | 6698 | hCoV-19/Australia/VIC5545/2020\|EPI_ISL_518374\|2020 | ···················· | ···················· | ···················· | ···················· | ···················· | ···················· | |
| G | 6699 | hCoV-19/Australia/VIC5546/2020\|EPI_ISL_518375\|2020 | ···················· | ···················· | ···················· | ···················· | ···················· | ···················· | |
| G | 6700 | hCoV-19/Australia/VIC5547/2020\|EPI_ISL_518376\|2020 | ···················· | ···················· | ···················· | ···················· | ···················· | ···················· | |

| | | | | | | | |
|---|---|---|---|---|---|---|---|
| G | 6701 hCoV-19/Australia/VIC5549/2020\|EPI_ISL_518378\|2020 | .................... | .................... | .................... | .................... | .................... | .................... |
| G | 6702 hCoV-19/Australia/VIC5552/2020\|EPI_ISL_518380\|2020 | .................... | .................... | .................... | .................... | .................... | .................... |
| G | 6703 hCoV-19/Australia/VIC5553/2020\|EPI_ISL_518381\|2020 | .................... | .................... | .................... | .................... | .................... | .................... |
| G | 6704 hCoV-19/Australia/VIC5554/2020\|EPI_ISL_518382\|2020 | .................... | .................... | .................... | .................... | .................... | .................... |
| G | 6705 hCoV-19/Australia/VIC5555/2020\|EPI_ISL_518383\|2020 | .................... | .................... | .................... | .................... | .................... | .................... |
| G | 6706 hCoV-19/Australia/VIC5556/2020\|EPI_ISL_518384\|2020 | .................... | .................... | .................... | .................... | .................... | .................... |
| G | 6707 hCoV-19/Australia/VIC5567/2020\|EPI_ISL_518386\|2020 | .................... | .................... | .................... | .................... | .................... | .................... |
| G | 6708 hCoV-19/Australia/VIC5568/2020\|EPI_ISL_518387\|2020 | .................... | .................... | .................... | .................... | .................... | .................... |
| G | 6709 hCoV-19/Australia/VIC5570/2020\|EPI_ISL_518389\|2020 | .................... | .................... | .................... | .................... | .................... | .................... |
| G | 6710 hCoV-19/Australia/VIC5572/2020\|EPI_ISL_518391\|2020 | .................... | .................... | .................... | .................... | .................... | .................... |
| G | 6711 hCoV-19/Australia/VIC5575/2020\|EPI_ISL_518393\|2020 | .................... | .................... | .................... | .................... | .................... | .................... |
| G | 6712 hCoV-19/Australia/VIC5576/2020\|EPI_ISL_518394\|2020 | .................... | .................... | .................... | .................... | .................... | .................... |
| G | 6713 hCoV-19/Australia/VIC5577/2020\|EPI_ISL_518395\|2020 | .................... | .................... | .................... | .................... | .................... | .................... |
| G | 6714 hCoV-19/Australia/VIC5578/2020\|EPI_ISL_518396\|2020 | .................... | .................... | .................... | .................... | .................... | .................... |
| G | 6715 hCoV-19/Australia/VIC5579/2020\|EPI_ISL_518397\|2020 | .................... | .................... | .................... | .................... | .................... | .................... |
| G | 6716 hCoV-19/Australia/VIC5580/2020\|EPI_ISL_518398\|2020 | .................... | .................... | .................... | .................... | .................... | .................... |
| G | 6717 hCoV-19/Australia/VIC5581/2020\|EPI_ISL_518399\|2020 | .................... | .................... | .................... | .................... | .................... | .................... |
| G | 6718 hCoV-19/United Arab Emirates/L3290220847/2020\|EPI_ | .................... | .................... | .................... | .................... | .................... | .................... |
| G | 6719 hCoV-19/United Arab Emirates/L3290222605/2020\|EPI_ | .................... | .................... | .................... | .................... | .................... | .................... |
| G | 6720 hCoV-19/United Arab Emirates/L3290223961/2020\|EPI_ | .................... | .................... | .................... | .................... | .................... | .................... |
| G | 6721 hCoV-19/United Arab Emirates/L3290224549/2020\|EPI_ | .................... | .................... | .................... | .................... | .................... | .................... |
| G | 6722 hCoV-19/United Arab Emirates/L3290235819/2020\|EPI_ | .................... | .................... | .................... | .................... | .................... | .................... |
| G | 6723 hCoV-19/United Arab Emirates/L3290238519/2020\|EPI_ | .................... | .................... | .................... | .................... | .................... | .................... |
| G | 6724 hCoV-19/United Arab Emirates/L3290248480/2020\|EPI_ | .................... | .................... | .................... | .................... | .................... | .................... |
| G | 6725 hCoV-19/United Arab Emirates/L3290248500/2020\|EPI_ | .................... | .................... | .................... | .................... | .................... | .................... |
| G | 6726 hCoV-19/United Arab Emirates/L3290250569/2020\|EPI_ | .................... | .................... | .................... | .................... | .................... | .................... |
| G | 6727 hCoV-19/United Arab Emirates/L3290250732/2020\|EPI_ | .................... | .................... | .................... | .................... | .................... | .................... |
| G | 6728 hCoV-19/United Arab Emirates/L3290250769/2020\|EPI_ | .................... | .................... | .................... | .................... | .................... | .................... |
| G | 6729 hCoV-19/United Arab Emirates/L3290250781/2020\|EPI_ | .................... | .................... | .................... | .................... | .................... | .................... |
| G | 6730 hCoV-19/USA/UT-08433/2020\|EPI_ISL_524209\|2020-05-2 | .................... | .................... | .................... | .................... | .................... | .................... |
| G | 6731 hCoV-19/USA/UT-08434/2020\|EPI_ISL_524210\|2020-05-1 | .................... | .................... | .................... | .................... | .........A.......... | .................... |
| G | 6732 hCoV-19/USA/UT-08439/2020\|EPI_ISL_524214\|2020-05-2 | .................... | .................... | .................... | .................... | .................... | .................... |
| G | 6733 hCoV-19/USA/UT-08440/2020\|EPI_ISL_524215\|2020-05-2 | .................... | .................... | .................... | .................... | .................... | .................... |
| G | 6734 hCoV-19/USA/UT-08441/2020\|EPI_ISL_524216\|2020-05-2 | .................... | .................... | .................... | .................... | .................... | .................... |
| G | 6735 hCoV-19/USA/UT-08442/2020\|EPI_ISL_524217\|2020-05-2 | .................... | .................... | .................... | .................... | .................... | .................... |
| G | 6736 hCoV-19/USA/UT-08446/2020\|EPI_ISL_524220\|2020-05-1 | .................... | .................... | .................... | .................... | .................... | .................... |
| G | 6737 hCoV-19/USA/UT-08452/2020\|EPI_ISL_524224\|2020-05-1 | .................... | .................... | .................... | .................... | .................... | .................... |
| G | 6738 hCoV-19/USA/UT-08453/2020\|EPI_ISL_524225\|2020-05-1 | .................... | .................... | .................... | .................... | .................... | .................... |
| G | 6739 hCoV-19/USA/UT-08454/2020\|EPI_ISL_524226\|2020-05-2 | .................... | .................... | .................... | .................... | .................... | .................... |
| G | 6740 hCoV-19/USA/UT-08455/2020\|EPI_ISL_524227\|2020-05-1 | .................... | .................... | .................... | .................... | .................... | .................... |
| G | 6741 hCoV-19/USA/UT-08456/2020\|EPI_ISL_524228\|2020-05-2 | .................... | .................... | .................... | .................... | .................... | .................... |
| G | 6742 hCoV-19/USA/UT-08459/2020\|EPI_ISL_524231\|2020-05-2 | .................... | .................... | .................... | .................... | .................... | .................... |
| G | 6743 hCoV-19/USA/UT-08460/2020\|EPI_ISL_524232\|2020-05-2 | .................... | .................... | .................... | .................... | .................... | .................... |
| G | 6744 hCoV-19/USA/UT-08464/2020\|EPI_ISL_524236\|2020-05-2 | .................... | .................... | .................... | .................... | .................... | .................... |
| G | 6745 hCoV-19/USA/UT-08466/2020\|EPI_ISL_524237\|2020-05-1 | .................... | .................... | .................... | .................... | .................... | .................... |
| G | 6746 hCoV-19/USA/UT-08468/2020\|EPI_ISL_524239\|2020-05-2 | .................... | .................... | .................... | .................... | .................... | .................... |
| G | 6747 hCoV-19/USA/UT-08469/2020\|EPI_ISL_524240\|2020-05-2 | .................... | .................... | .................... | .................... | .................... | .................... |
| G | 6748 hCoV-19/USA/UT-08471/2020\|EPI_ISL_524242\|2020-05-2 | .................... | .................... | .................... | .................... | .................... | .................... |
| G | 6749 hCoV-19/USA/UT-08475/2020\|EPI_ISL_524245\|2020-05-2 | .................... | .................... | .................... | .................... | .................... | .................... |
| G | 6750 hCoV-19/USA/UT-08477/2020\|EPI_ISL_524247\|2020-05-1 | .................... | .................... | .................... | .................... | .................... | .................... |
| G | 6751 hCoV-19/USA/UT-08480/2020\|EPI_ISL_524249\|2020-05-2 | .................... | .................... | .................... | .................... | .................... | .................... |
| G | 6752 hCoV-19/France/ARA-10259/2020\|EPI_ISL_525539\|2020- | .................... | .................... | .................... | .................... | .................... | .................... |
| G | 6753 hCoV-19/France/ARA-10836/2020\|EPI_ISL_525540\|2020- | .................... | .................... | .................... | .................... | .................... | .................... |
| G | 6754 hCoV-19/France/ARA-11167/2020\|EPI_ISL_525541\|2020- | .................... | .................... | .................... | .................... | .................... | .................... |
| G | 6755 hCoV-19/France/ARA-11228/2020\|EPI_ISL_525542\|2020- | .................... | .................... | .................... | .................... | .................... | .................... |
| G | 6756 hCoV-19/France/ARA-11231/2020\|EPI_ISL_525543\|2020- | .................... | .................... | .................... | .................... | .................... | .................... |
| G | 6757 hCoV-19/Italy/APU-UniMI-102/2020\|EPI_ISL_525553\|20 | .................... | .................... | .................... | .................... | .................... | .................... |
| G | 6758 hCoV-19/Italy/APU-UniMI-368/2020\|EPI_ISL_525557\|20 | .................... | .................... | .................... | .................... | .................... | .................... |
| G | 6759 hCoV-19/Italy/APU-UniMI-5409/2020\|EPI_ISL_525560\|2 | .................... | .................... | .................... | ..............A..... | .................... | .................... |
| G | 6760 hCoV-19/Italy/APU-UniMI-5410/2020\|EPI_ISL_525561\|2 | .................... | .................... | .................... | .................... | .................... | .................... |
| G | 6761 hCoV-19/Italy/APU-UniMI-5439/2020\|EPI_ISL_525569\|2 | .................... | .................... | .................... | .................... | .................... | .................... |
| G | 6762 hCoV-19/Italy/APU-UniMI-6/2020\|EPI_ISL_525570\|2020 | .................... | .................... | .................... | .................... | .................... | .................... |
| G | 6763 hCoV-19/Italy/BAS-UniMI-2956/2020\|EPI_ISL_525573\|2 | .................... | .................... | .................... | .................... | .................... | .................... |
| G | 6764 hCoV-19/Italy/BAS-UniMI-4160/2020\|EPI_ISL_525574\|2 | .................... | .................... | .................... | .................... | .................... | .................... |
| G | 6765 hCoV-19/USA/NY-Wadsworth-10131-01/2020\|EPI_ISL_525 | .................... | .................... | .................... | .................... | .................... | .................... |
| G | 6766 hCoV-19/USA/NY-Wadsworth-10713-01/2020\|EPI_ISL_525 | .................... | .................... | .................... | .................... | .................... | .................... |
| G | 6767 hCoV-19/USA/NY-Wadsworth-10715-01/2020\|EPI_ISL_525 | .................... | .................... | .................... | .................... | .................... | .................... |
| G | 6768 hCoV-19/USA/NY-Wadsworth-10715-2/2020\|EPI_ISL_5255 | .................... | .................... | .................... | .................... | .................... | .................... |
| G | 6769 hCoV-19/USA/NY-Wadsworth-11171-01/2020\|EPI_ISL_525 | .................... | .................... | .................... | .................... | .................... | .................... |
| G | 6770 hCoV-19/USA/VA-DCLS-0579/2020\|EPI_ISL_526800\|2020- | .................... | .................... | .................... | .................... | .................... | .................... |
| G | 6771 hCoV-19/USA/VA-DCLS-0580/2020\|EPI_ISL_526801\|2020- | .................... | .................... | .................... | .................... | .................... | .................... |
| G | 6772 hCoV-19/USA/VA-DCLS-0581/2020\|EPI_ISL_526802\|2020- | .................... | .................... | .................... | .................... | .................... | .................... |
| G | 6773 hCoV-19/USA/VA-DCLS-0582/2020\|EPI_ISL_526803\|2020- | .................... | .................... | .................... | .................... | .................... | .................... |
| G | 6774 hCoV-19/USA/VA-DCLS-0583/2020\|EPI_ISL_526804\|2020- | .................... | .................... | .................... | .................... | .................... | .................... |
| G | 6775 hCoV-19/USA/VA-DCLS-0584/2020\|EPI_ISL_526805\|2020- | .................... | .................... | .................... | .................... | .................... | .................... |
| G | 6776 hCoV-19/USA/VA-DCLS-0585/2020\|EPI_ISL_526806\|2020- | .................... | .................... | .................... | .................... | .................... | .................... |
| G | 6778 hCoV-19/USA/VA-DCLS-0588/2020\|EPI_ISL_526808\|2020- | .................... | .................... | .................... | .................... | .................... | .................... |
| G | 6779 hCoV-19/USA/VA-DCLS-0589/2020\|EPI_ISL_526809\|2020- | .................... | .................... | .................... | .................... | .................... | .................... |
| G | 6780 hCoV-19/USA/VA-DCLS-0590/2020\|EPI_ISL_526810\|2020- | .................... | .................... | .................... | .................... | .................... | .................... |
| G | 6781 hCoV-19/USA/VA-DCLS-0591/2020\|EPI_ISL_526811\|2020- | .................... | .................... | .................... | .................... | .................... | .................... |

| | | | | | | | | |
|---|---|---|---|---|---|---|---|---|
| G | 6782 | hCoV-19/USA/VA-DCLS-0592/2020\|EPI_ISL_526812\|2020- | ···················· | ···················· | ···················· | ···················· | ···················· | ···················· |
| G | 6783 | hCoV-19/USA/VA-DCLS-0593/2020\|EPI_ISL_526813\|2020- | ···················· | ···················· | ···················· | ···················· | ···················· | ···················· |
| G | 6784 | hCoV-19/USA/VA-DCLS-0594/2020\|EPI_ISL_526814\|2020- | ···················· | ···················· | ···················· | ···················· | ···················· | ···················· |
| G | 6785 | hCoV-19/USA/VA-DCLS-0595/2020\|EPI_ISL_526815\|2020- | ···················· | ···················· | ···················· | ···················· | ···················· | ···················· |
| G | 6786 | hCoV-19/USA/VA-DCLS-0596/2020\|EPI_ISL_526816\|2020- | ···················· | ···················· | ···················· | ···················· | ···················· | ···················· |
| G | 6787 | hCoV-19/USA/VA-DCLS-0599/2020\|EPI_ISL_526817\|2020- | ···················· | ···················· | ···················· | ···················· | ···················· | ···················· |
| G | 6788 | hCoV-19/USA/VA-DCLS-0600/2020\|EPI_ISL_526818\|2020- | ···················· | ···················· | ···················· | ···················· | ···················· | ···················· |
| G | 6789 | hCoV-19/USA/VA-DCLS-0603/2020\|EPI_ISL_526820\|2020- | ···················· | ···················· | ···················· | ···················· | ···················· | ···················· |
| G | 6790 | hCoV-19/USA/VA-DCLS-0604/2020\|EPI_ISL_526821\|2020- | ···················· | ···················· | ···················· | ···················· | ···················· | ···················· |
| G | 6791 | hCoV-19/USA/VA-DCLS-0606/2020\|EPI_ISL_526823\|2020- | ···················· | ···················· | ···················· | ···················· | ···················· | ···················· |
| G | 6792 | hCoV-19/USA/VA-DCLS-0607/2020\|EPI_ISL_526824\|2020- | ···················· | ···················· | ···················· | ···················· | ···················· | ···················· |
| G | 6793 | hCoV-19/USA/VA-DCLS-0753/2020\|EPI_ISL_526869\|2020- | ···················· | ···················· | ···················· | ···················· | ···················· | ···················· |
| G | 6794 | hCoV-19/USA/VA-DCLS-0754/2020\|EPI_ISL_526870\|2020- | ···················· | ···················· | ···················· | ···················· | ···················· | ···················· |
| G | 6795 | hCoV-19/USA/VA-DCLS-0755/2020\|EPI_ISL_526871\|2020- | ···················· | ···················· | ···················· | ···················· | ···················· | ···················· |
| G | 6796 | hCoV-19/USA/VA-DCLS-0756/2020\|EPI_ISL_526872\|2020- | ···················· | ···················· | ···················· | ···················· | ···················· | ···················· |
| G | 6797 | hCoV-19/USA/VA-DCLS-0757/2020\|EPI_ISL_526873\|2020- | ···················· | ···················· | ···················· | ···················· | ···················· | ···················· |
| G | 6798 | hCoV-19/USA/VA-DCLS-0758/2020\|EPI_ISL_526874\|2020- | ···················· | ···················· | ···················· | ···················· | ···················· | ···················· |
| G | 6799 | hCoV-19/USA/VA-DCLS-0759/2020\|EPI_ISL_526875\|2020- | ···················· | ···················· | ···················· | ···················· | ···················· | ···················· |
| G | 6800 | hCoV-19/USA/VA-DCLS-0760/2020\|EPI_ISL_526876\|2020- | ···················· | ···················· | ···················· | ···················· | ···················· | ···················· |
| G | 6801 | hCoV-19/USA/VA-DCLS-0761/2020\|EPI_ISL_526877\|2020- | ···················· | ···················· | ···················· | ···················· | ···················· | ···················· |
| G | 6802 | hCoV-19/USA/VA-DCLS-0762/2020\|EPI_ISL_526878\|2020- | ···················· | ···················· | ···················· | ···················· | ···················· | ···················· |
| G | 6803 | hCoV-19/USA/VA-DCLS-0763/2020\|EPI_ISL_526879\|2020- | ···················· | ···················· | ···················· | ···················· | ···················· | ···················· |
| G | 6804 | hCoV-19/USA/VA-DCLS-0764/2020\|EPI_ISL_526880\|2020- | ···················· | ···················· | ···················· | ···················· | ···················· | ···················· |
| G | 6805 | hCoV-19/USA/VA-DCLS-0765/2020\|EPI_ISL_526881\|2020- | ···················· | ···················· | ···················· | ···················· | ···················· | ···················· |
| G | 6806 | hCoV-19/USA/VA-DCLS-0766/2020\|EPI_ISL_526882\|2020- | ···················· | ···················· | ···················· | ···················· | ···················· | ···················· |
| G | 6807 | hCoV-19/USA/VA-DCLS-0767/2020\|EPI_ISL_526883\|2020- | ···················· | ···················· | ···················· | ···················· | ···················· | ···················· |
| G | 6808 | hCoV-19/USA/VA-DCLS-0768/2020\|EPI_ISL_526884\|2020- | ···················· | ···················· | ···················· | ···················· | ···················· | ···················· |
| G | 6809 | hCoV-19/USA/VA-DCLS-0770/2020\|EPI_ISL_526885\|2020- | ···················· | ···················· | ···················· | ···················· | ···················· | ···················· |
| G | 6810 | hCoV-19/USA/VA-DCLS-0771/2020\|EPI_ISL_526886\|2020- | ···················· | ···················· | ···················· | ···················· | ···················· | ···················· |
| G | 6811 | hCoV-19/USA/VA-DCLS-0772/2020\|EPI_ISL_526887\|2020- | ···················· | ···················· | ···················· | ···················· | ···················· | ···················· |
| G | 6812 | hCoV-19/USA/VA-DCLS-0773/2020\|EPI_ISL_526888\|2020- | ···················· | ···················· | ···················· | ···················· | ···················· | ···················· |
| G | 6813 | hCoV-19/USA/VA-DCLS-0775/2020\|EPI_ISL_526889\|2020- | ···················· | ···················· | ···················· | ···················· | ···················· | ···················· |
| G | 6814 | hCoV-19/USA/VA-DCLS-0776/2020\|EPI_ISL_526890\|2020- | ···················· | ···················· | ···················· | ···················· | ···················· | ···················· |
| G | 6815 | hCoV-19/USA/VA-DCLS-0777/2020\|EPI_ISL_526891\|2020- | ···················· | ···················· | ···················· | ···················· | ···················· | ···················· |
| G | 6816 | hCoV-19/USA/VA-DCLS-0778/2020\|EPI_ISL_526892\|2020- | ···················· | ···················· | ···················· | ···················· | ···················· | ···················· |
| G | 6817 | hCoV-19/USA/VA-DCLS-0779/2020\|EPI_ISL_526893\|2020- | ···················· | ···················· | ···················· | ···················· | ···················· | ···················· |
| G | 6818 | hCoV-19/USA/VA-DCLS-0780/2020\|EPI_ISL_526894\|2020- | ···················· | ···················· | ···················· | ···················· | ···················· | ···················· |
| G | 6819 | hCoV-19/USA/VA-DCLS-0790/2020\|EPI_ISL_526895\|2020- | ···················· | ···················· | ···················· | ···················· | ···················· | ···················· |
| G | 6820 | hCoV-19/USA/VA-DCLS-0791/2020\|EPI_ISL_526896\|2020- | ···················· | ···················· | ···················· | ···················· | ···················· | ···················· |
| G | 6821 | hCoV-19/USA/VA-DCLS-0793/2020\|EPI_ISL_526897\|2020- | ···················· | ···················· | ···················· | ···················· | ···················· | ···················· |
| G | 6822 | hCoV-19/USA/VA-DCLS-0794/2020\|EPI_ISL_526898\|2020- | ···················· | ···················· | ···················· | ···················· | ···················· | ···················· |
| G | 6823 | hCoV-19/USA/VA-DCLS-0997/2020\|EPI_ISL_526899\|2020- | ···················· | ···················· | ···················· | ···················· | ···················· | ···················· |
| G | 6824 | hCoV-19/Italy/ABR-IZSGC-TE28268/2020\|EPI_ISL_52902 | ···················· | ···················· | ···················· | ···················· | ···················· | ···················· |
| G | 6825 | hCoV-19/Italy/ABR-IZSGC-TE30364/2020\|EPI_ISL_52902 | ···················· | ···················· | ···················· | ···················· | ···················· | ···················· |
| G | 6826 | hCoV-19/Italy/ABR-IZSGC-TE36500/2020\|EPI_ISL_52902 | ···················· | ···················· | ···················· | ···················· | ···················· | ···················· |
| G | 6827 | hCoV-19/Italy/ABR-IZSGC-TE8387/2020\|EPI_ISL_529026 | ···················· | ···················· | ···················· | ···················· | ···················· | ···················· |
| G | 6828 | hCoV-19/England/20358026204/2020\|EPI_ISL_529028\|20 | ···················· | ···················· | ···················· | ···················· | ···················· | ···················· |
| G | 6829 | hCoV-19/Egypt/C-CU001a-S1/2020\|EPI_ISL_529031\|2020 | ···················· | ···················· | ···················· | ···················· | ···················· | ···················· |
| G | 6830 | hCoV-19/Egypt/C-CU002b-S3/2020\|EPI_ISL_529032\|2020 | ···················· | ···················· | ···················· | ···················· | ···················· | ···················· |
| G | 6831 | hCoV-19/USA/NY-Wadsworth-11460-01/2020\|EPI_ISL_529 | ···················· | ···················· | ···················· | ···················· | ···················· | ···················· |
| G | 6832 | hCoV-19/USA/NY-Wadsworth-11661-01/2020\|EPI_ISL_529 | ···················· | ···················· | ···················· | ···················· | ···················· | ···················· |
| G | 6833 | hCoV-19/USA/NY-WC-46316-01/2020\|EPI_ISL_529035\|202 | ···················· | ···················· | ···················· | ···················· | ···················· | ···················· |
| G | 6834 | hCoV-19/USA/NY-WC-46317-01/2020\|EPI_ISL_529036\|202 | ···················· | ···················· | ···················· | ···················· | ···················· | ···················· |
| G | 6835 | hCoV-19/Peru/LIM-UPCH-0003/2020\|EPI_ISL_529065\|202 | ···················· | ···················· | ···················· | ···················· | ···················· | ···················· |
| G | 6836 | hCoV-19/Peru/LIM-UPCH-0004/2020\|EPI_ISL_529066\|202 | ···················· | ···················· | ···················· | ···················· | ···················· | ···················· |
| G | 6837 | hCoV-19/Peru/LAM-UPCH-0006/2020\|EPI_ISL_529067\|202 | ···················· | ···················· | ···················· | ···················· | ···················· | ···················· |
| G | 6838 | hCoV-19/Peru/LIM-UPCH-0011/2020\|EPI_ISL_529068\|202 | ···················· | ···················· | ···················· | ···················· | ···················· | ···················· |
| G | 6839 | hCoV-19/Peru/LIM-UPCH-0012/2020\|EPI_ISL_529069\|202 | ···················· | ···················· | ···················· | ···················· | ···················· | ···················· |
| G | 6840 | hCoV-19/Peru/CAL-UPCH-0013/2020\|EPI_ISL_529070\|202 | ···················· | ···················· | ···················· | ···················· | ···················· | ···················· |
| G | 6841 | hCoV-19/Peru/LIM-UPCH-0014/2020\|EPI_ISL_529071\|202 | ···················· | ···················· | ···················· | ···················· | ···················· | ···················· |
| G | 6842 | hCoV-19/Peru/LIM-UPCH-0022/2020\|EPI_ISL_529072\|202 | ···················· | ···················· | ···················· | ···················· | ···················· | ···················· |
| G | 6843 | hCoV-19/Peru/CAL-UPCH-0036/2020\|EPI_ISL_529073\|202 | ···················· | ···················· | ···················· | ···················· | ···················· | ···················· |
| G | 6844 | hCoV-19/Peru/CAL-UPCH-0037/2020\|EPI_ISL_529074\|202 | ···················· | ···················· | ···················· | ···················· | ···················· | ···················· |
| G | 6845 | hCoV-19/Peru/LIM-UPCH-0038/2020\|EPI_ISL_529075\|202 | ···················· | ···················· | ···················· | ···················· | ···················· | ···················· |
| G | 6846 | hCoV-19/Peru/LIM-UPCH-0039/2020\|EPI_ISL_529076\|202 | ···················· | ···················· | ···················· | ···················· | ···················· | ···················· |
| G | 6847 | hCoV-19/Peru/LIM-UPCH-0041/2020\|EPI_ISL_529077\|202 | ···················· | ···················· | ···················· | ···················· | ···················· | ···················· |
| G | 6848 | hCoV-19/Peru/ANC-UPCH-0043/2020\|EPI_ISL_529078\|202 | ···················· | ···················· | ···················· | ···················· | ···················· | ···················· |
| G | 6849 | hCoV-19/Peru/CAL-UPCH-0044/2020\|EPI_ISL_529079\|202 | ···················· | ···················· | ···················· | ···················· | ···················· | ···················· |
| G | 6850 | hCoV-19/USA/SC-DHEC-0515/2020\|EPI_ISL_529080\|2020- | ···················· | ···················· | ···················· | ···················· | ···················· | ···················· |
| G | 6851 | hCoV-19/USA/SC-DHEC-0533/2020\|EPI_ISL_529081\|2020- | ···················· | ···················· | ···················· | ···················· | ···················· | ···················· |
| G | 6852 | hCoV-19/USA/SC-DHEC-0534/2020\|EPI_ISL_529082\|2020- | ···················· | ···················· | ···················· | ···················· | ···················· | ···················· |
| G | 6854 | hCoV-19/USA/SC-DHEC-0537/2020\|EPI_ISL_529084\|2020- | ···················· | ···················· | ···················· | ···················· | ···················· | ···················· |
| G | 6855 | hCoV-19/USA/SC-DHEC-0545/2020\|EPI_ISL_529085\|2020- | ···················· | ···················· | ···················· | ···················· | ···················· | ···················· |
| G | 6856 | hCoV-19/USA/SC-DHEC-0546/2020\|EPI_ISL_529086\|2020- | ···················· | ···················· | ···················· | ···················· | ···················· | ···················· |
| G | 6857 | hCoV-19/USA/SC-DHEC-0548/2020\|EPI_ISL_529087\|2020- | ···················· | ···················· | ···················· | ···················· | ···················· | ···················· |
| G | 6858 | hCoV-19/USA/SC-DHEC-0549/2020\|EPI_ISL_529088\|2020- | ···················· | ···················· | ···················· | ···················· | ···················· | ···················· |
| G | 6859 | hCoV-19/USA/SC-DHEC-0550/2020\|EPI_ISL_529089\|2020- | ···················· | ···················· | ···················· | ···················· | ···················· | ···················· |
| G | 6860 | hCoV-19/USA/SC-DHEC-0551/2020\|EPI_ISL_529090\|2020- | ···················· | ···················· | ···················· | ···················· | ···················· | ···················· |
| G | 6861 | hCoV-19/USA/SC-DHEC-0576/2020\|EPI_ISL_529091\|2020- | ···················· | ·········A·········· | ···················· | ···················· | ···················· | ···················· |
| G | 6862 | hCoV-19/USA/SC-DHEC-0583/2020\|EPI_ISL_529092\|2020- | ···················· | ···················· | ···················· | ···················· | ···················· | ···················· |

| | | | | | | | | |
|---|---|---|---|---|---|---|---|---|
| G | 6863 | hCoV-19/USA/SC-DHEC-0584/2020\|EPI_ISL_529093\|2020- | ···················· | ···················· | ···················· | ···················· | ···················· | ···················· |
| G | 6864 | hCoV-19/USA/SC-DHEC-0585/2020\|EPI_ISL_529094\|2020- | ···················· | ···················· | ···················· | ···················· | ···················· | ···················· |
| G | 6865 | hCoV-19/USA/SC-DHEC-0586/2020\|EPI_ISL_529095\|2020- | ···················· | ···················· | ···················· | ···················· | ···················· | ···················· |
| G | 6866 | hCoV-19/USA/SC-DHEC-0587/2020\|EPI_ISL_529096\|2020- | ···················· | ···················· | ···················· | ···················· | ···················· | ···················· |
| G | 6867 | hCoV-19/Spain/MD-H12-73/2020\|EPI_ISL_530004\|2020-0 | ···················· | ···················· | ···················· | ···················· | ···················· | ···················· |
| G | 6868 | hCoV-19/Spain/MD-H12-75/2020\|EPI_ISL_530005\|2020-0 | ···················· | ···················· | ···················· | ···················· | ···················· | ···················· |
| G | 6869 | hCoV-19/Spain/MD-H12-76/2020\|EPI_ISL_530006\|2020-0 | ···················· | ···················· | ···················· | ···················· | ···················· | ···················· |
| G | 6870 | hCoV-19/Spain/MD-H12-77/2020\|EPI_ISL_530007\|2020-0 | ···················· | ···················· | ···················· | ···················· | ···················· | ···A················ |
| G | 6871 | hCoV-19/Spain/MD-H12-78/2020\|EPI_ISL_530008\|2020-0 | ···················· | ···················· | ···················· | ···················· | ···················· | ···················· |
| G | 6872 | hCoV-19/Spain/MD-H12-80/2020\|EPI_ISL_530009\|2020-0 | ···················· | ···················· | ···················· | ···················· | ···················· | ···················· |
| G | 6873 | hCoV-19/Spain/MD-H12-81/2020\|EPI_ISL_530010\|2020-0 | ···················· | ···················· | ···················· | ···················· | ···················· | ···················· |
| G | 6874 | hCoV-19/Spain/MD-H12-82/2020\|EPI_ISL_530011\|2020-0 | ···················· | ···················· | ···················· | ···················· | ···················· | ···················· |
| G | 6875 | hCoV-19/Spain/MD-H12-83/2020\|EPI_ISL_530012\|2020-0 | ···················· | ···················· | ···················· | ···················· | ···················· | ···················· |
| G | 6876 | hCoV-19/Spain/MD-H12-84/2020\|EPI_ISL_530013\|2020-0 | ···················· | ···················· | ···················· | ···················· | ···················· | ···················· |
| G | 6877 | hCoV-19/Spain/MD-H12-85/2020\|EPI_ISL_530014\|2020-0 | ···················· | ···················· | ···················· | ···················· | ···················· | ···················· |
| G | 6878 | hCoV-19/Spain/MD-H12-87/2020\|EPI_ISL_530015\|2020-0 | ···················· | ·A·················· | ···················· | ···················· | ···················· | ···················· |
| G | 6879 | hCoV-19/Spain/MD-H12-89/2020\|EPI_ISL_530016\|2020-0 | ···················· | ···················· | ···················· | ···················· | ···················· | ···················· |
| G | 6880 | hCoV-19/Spain/MD-H12-90/2020\|EPI_ISL_530017\|2020-0 | ···················· | ···················· | ···················· | ···················· | ···················· | ···················· |
| G | 6881 | hCoV-19/Spain/MD-H12-91/2020\|EPI_ISL_530018\|2020-0 | ···················· | ···················· | ···················· | ···················· | ···················· | ···················· |
| G | 6882 | hCoV-19/Spain/MD-H12-93/2020\|EPI_ISL_530019\|2020-0 | ···················· | ···················· | ···················· | ···················· | ···················· | ···················· |
| G | 6883 | hCoV-19/Spain/MD-H12-94/2020\|EPI_ISL_530020\|2020-0 | ···················· | ···················· | ···················· | ···················· | ···················· | ···················· |
| G | 6884 | hCoV-19/Spain/MD-H12-95/2020\|EPI_ISL_530021\|2020-0 | ···················· | ···················· | ···················· | ···················· | ···················· | ···················· |
| G | 6885 | hCoV-19/Spain/MD-H12-96/2020\|EPI_ISL_530022\|2020-0 | ···················· | ···················· | ···················· | ···················· | ···················· | ···················· |
| G | 6886 | hCoV-19/Spain/MD-H12-97/2020\|EPI_ISL_530023\|2020-0 | ···················· | ···················· | ···················· | ···················· | ···················· | ···················· |
| G | 6887 | hCoV-19/Spain/MD-H12-98/2020\|EPI_ISL_530024\|2020-0 | ···················· | ···················· | ···················· | ···················· | ···················· | ···················· |
| G | 6888 | hCoV-19/Spain/MD-HLP-100-1/2020\|EPI_ISL_530025\|202 | ···················· | ···················· | ···················· | ···················· | ···················· | ···················· |
| G | 6889 | hCoV-19/Spain/MD-HLP-101/2020\|EPI_ISL_530027\|2020- | ···················· | ···················· | ···················· | ···················· | ···················· | ···················· |
| G | 6890 | hCoV-19/Spain/MD-HLP-102-1/2020\|EPI_ISL_530028\|202 | ···················· | ···················· | ···················· | ···················· | ···················· | ···················· |
| G | 6891 | hCoV-19/Spain/MD-HLP-102-2/2020\|EPI_ISL_530029\|202 | ···················· | ···················· | ···················· | ···················· | ···················· | ···················· |
| G | 6892 | hCoV-19/Spain/MD-HLP-103/2020\|EPI_ISL_530030\|2020- | ···················· | ···················· | ···················· | ···················· | ···················· | ···················· |
| G | 6893 | hCoV-19/Spain/MD-HLP-104-1/2020\|EPI_ISL_530031\|202 | ···················· | ···················· | ···················· | ···················· | ···················· | ···················· |
| G | 6894 | hCoV-19/Spain/MD-HLP-104-2/2020\|EPI_ISL_530032\|202 | ···················· | ···················· | ···················· | ···················· | ···················· | ···················· |
| G | 6895 | hCoV-19/Spain/MD-HLP-104-3/2020\|EPI_ISL_530033\|202 | ···················· | ···················· | ···················· | ···················· | ···················· | ···················· |
| G | 6896 | hCoV-19/Spain/MD-HLP-106-1/2020\|EPI_ISL_530034\|202 | ···················· | ···················· | ···················· | ···················· | ···················· | ···················· |
| G | 6897 | hCoV-19/Spain/MD-HLP-106-2/2020\|EPI_ISL_530035\|202 | ···················· | ···················· | ···················· | ···················· | ···················· | ···················· |
| G | 6898 | hCoV-19/Spain/MD-HLP-106-3/2020\|EPI_ISL_530036\|202 | ···················· | ···················· | ···················· | ···················· | ···················· | ···················· |
| G | 6899 | hCoV-19/Spain/MD-HLP-108/2020\|EPI_ISL_530037\|2020- | ···················· | ···················· | ···················· | ···················· | ···················· | ···················· |
| G | 6900 | hCoV-19/Spain/MD-HLP-109/2020\|EPI_ISL_530038\|2020- | ···················· | ···················· | ···················· | ···················· | ···················· | ···················· |
| G | 6901 | hCoV-19/Spain/MD-HLP-110/2020\|EPI_ISL_530039\|2020- | ···················· | ···················· | ···················· | ···················· | ···················· | ···················· |
| G | 6902 | hCoV-19/Spain/MD-HLP-111/2020\|EPI_ISL_530040\|2020- | ···················· | ···················· | ···················· | ···················· | ···················· | ···················· |
| G | 6903 | hCoV-19/Spain/MD-HLP-94/2020\|EPI_ISL_530090\|2020-0 | ···················· | ···················· | ···················· | ···················· | ···················· | ···················· |
| G | 6904 | hCoV-19/Spain/MD-HLP-95/2020\|EPI_ISL_530091\|2020-0 | ···················· | ···················· | ···················· | ···················· | ···················· | ···················· |
| G | 6905 | hCoV-19/Spain/MD-HLP-96/2020\|EPI_ISL_530092\|2020-0 | ···················· | ···················· | ···················· | ···················· | ···················· | ···················· |
| G | 6906 | hCoV-19/Spain/MD-HLP-97/2020\|EPI_ISL_530093\|2020-0 | ···················· | ···················· | ···················· | ···················· | ···················· | ···················· |
| G | 6907 | hCoV-19/Spain/MD-H12-R11/2020\|EPI_ISL_530094\|2020- | ···················· | ···················· | ···················· | ···················· | ···················· | ···················· |
| G | 6908 | hCoV-19/Spain/MD-H12-R17/2020\|EPI_ISL_530095\|2020- | ···················· | ···················· | ···················· | ···················· | ···················· | ···················· |
| G | 6909 | hCoV-19/Spain/MD-H12-R19/2020\|EPI_ISL_530096\|2020- | ···················· | ···················· | ···················· | ···················· | ···················· | ···················· |
| G | 6910 | hCoV-19/Spain/MD-H12-R20/2020\|EPI_ISL_530097\|2020- | ···················· | ···················· | ···················· | ···················· | ···················· | ···················· |
| G | 6911 | hCoV-19/Spain/MD-H12-R22/2020\|EPI_ISL_530098\|2020- | ···················· | ···················· | ···················· | ···················· | ···················· | ···················· |
| G | 6912 | hCoV-19/Spain/MD-H12-R25/2020\|EPI_ISL_530099\|2020- | ···················· | ···················· | ···················· | ···················· | ···················· | ···················· |
| G | 6913 | hCoV-19/England/LIVE-A837A/2020\|EPI_ISL_517151\|202 | ···················· | ···················· | ···················· | ···················· | ···················· | ···················· |
| G | 6914 | hCoV-19/England/LIVE-A83B6/2020\|EPI_ISL_517155\|202 | ···················· | ···················· | ···················· | ···················· | ···················· | ···················· |
| G | 6915 | hCoV-19/England/LIVE-A83F2/2020\|EPI_ISL_517159\|202 | ···················· | ···················· | ···················· | ···················· | ···················· | ···················· |
| G | 6916 | hCoV-19/Australia/VIC5583/2020\|EPI_ISL_518401\|2020 | ···················· | ···················· | ···················· | ···················· | ···················· | ···················· |
| G | 6918 | hCoV-19/Australia/VIC5586/2020\|EPI_ISL_518403\|2020 | ···················· | ···················· | ···················· | ···················· | ···················· | ···················· |
| G | 6919 | hCoV-19/Australia/VIC5587/2020\|EPI_ISL_518404\|2020 | ···················· | ···················· | ···················· | ···················· | ···················· | ···················· |
| G | 6920 | hCoV-19/Australia/VIC5588/2020\|EPI_ISL_518405\|2020 | ···················· | ···················· | ···················· | ···················· | ···················· | ···················· |
| G | 6921 | hCoV-19/Australia/VIC5589/2020\|EPI_ISL_518406\|2020 | ···················· | ···················· | ···················· | ···················· | ···················· | ···················· |
| G | 6922 | hCoV-19/Australia/VIC5590/2020\|EPI_ISL_518407\|2020 | ···················· | ···················· | ···················· | ···················· | ···················· | ···················· |
| G | 6923 | hCoV-19/Australia/VIC5673/2020\|EPI_ISL_518480\|2020 | ···················· | ···················· | ···················· | ···················· | ···················· | ···················· |
| G | 6924 | hCoV-19/Australia/VIC5675/2020\|EPI_ISL_518482\|2020 | ···················· | ···················· | ···················· | ···················· | ···················· | ···················· |
| G | 6925 | hCoV-19/Australia/VIC5676/2020\|EPI_ISL_518483\|2020 | ···················· | ···················· | ···················· | ···················· | ···················· | ···················· |
| G | 6926 | hCoV-19/Australia/VIC5677/2020\|EPI_ISL_518484\|2020 | ···················· | ···················· | ···················· | ···················· | ···················· | ···················· |
| G | 6927 | hCoV-19/Australia/VIC5678/2020\|EPI_ISL_518485\|2020 | ···················· | ···················· | ···················· | ···················· | ···················· | ···················· |
| G | 6928 | hCoV-19/Australia/VIC5679/2020\|EPI_ISL_518486\|2020 | ···················· | ···················· | ···················· | ···················· | ···················· | ···················· |
| G | 6929 | hCoV-19/Australia/VIC5680/2020\|EPI_ISL_518487\|2020 | ···················· | ···················· | ···················· | ···················· | ···················· | ···················· |
| G | 6930 | hCoV-19/Australia/VIC5682/2020\|EPI_ISL_518488\|2020 | ···················· | ···················· | ···················· | ···················· | ···················· | ···················· |
| G | 6931 | hCoV-19/Australia/VIC4404/2020\|EPI_ISL_519700\|2020 | ···················· | ···················· | ···················· | ···················· | ···················· | ···················· |
| G | 6932 | hCoV-19/Australia/VIC4405/2020\|EPI_ISL_519701\|2020 | ···················· | ···················· | ···················· | ···················· | ···················· | ···················· |
| G | 6933 | hCoV-19/Australia/VIC4406/2020\|EPI_ISL_519702\|2020 | ···················· | ···················· | ···················· | ···················· | ···················· | ···················· |
| G | 6934 | hCoV-19/Australia/VIC4407/2020\|EPI_ISL_519703\|2020 | ···················· | ···················· | ···················· | ···················· | ···················· | ···················· |
| G | 6935 | hCoV-19/Australia/VIC4408/2020\|EPI_ISL_519704\|2020 | ···················· | ···················· | ···················· | ···················· | ···················· | ···················· |
| G | 6936 | hCoV-19/Australia/VIC4409/2020\|EPI_ISL_519705\|2020 | ···················· | ···················· | ···················· | ···················· | ···················· | ···················· |
| G | 6937 | hCoV-19/Australia/VIC4410/2020\|EPI_ISL_519706\|2020 | ···················· | ···················· | ···················· | ···················· | ···················· | ···················· |
| G | 6938 | hCoV-19/Australia/VIC4411/2020\|EPI_ISL_519707\|2020 | ···················· | ···················· | ···················· | ···················· | ···················· | ···················· |
| G | 6939 | hCoV-19/Australia/VIC4412/2020\|EPI_ISL_519708\|2020 | ···················· | ···················· | ···················· | ···················· | ···················· | ···················· |
| G | 6940 | hCoV-19/Australia/VIC4413/2020\|EPI_ISL_519709\|2020 | ···················· | ···················· | ···················· | ···················· | ···················· | ···················· |
| G | 6941 | hCoV-19/Australia/VIC4414/2020\|EPI_ISL_519710\|2020 | ···················· | ···················· | ···················· | ···················· | ···················· | ···················· |
| G | 6942 | hCoV-19/Australia/VIC4415/2020\|EPI_ISL_519711\|2020 | ···················· | ···················· | ···················· | ···················· | ···················· | ···················· |
| G | 6943 | hCoV-19/Australia/VIC4416/2020\|EPI_ISL_519712\|2020 | ···················· | ···················· | ···················· | ···················· | ···················· | ···················· |

| | | | | | | | | | |
|---|---|---|---|---|---|---|---|---|---|
| G | 6944 | hCoV-19/Australia/VIC4417/2020\|EPI_ISL_519713\|2020 | ···················· | ···················· | ···················· | ···················· | ···················· | ···················· | |
| G | 6945 | hCoV-19/Australia/VIC4418/2020\|EPI_ISL_519714\|2020 | ···················· | ···················· | ···················· | ···················· | ···················· | ···················· | |
| G | 6946 | hCoV-19/Australia/VIC4419/2020\|EPI_ISL_519715\|2020 | ···················· | ···················· | ···················· | ···················· | ···················· | ···················· | |
| G | 6947 | hCoV-19/Australia/VIC4420/2020\|EPI_ISL_519716\|2020 | ···················· | ···················· | ···················· | ···················· | ···················· | ···················· | |
| G | 6948 | hCoV-19/Australia/VIC4421/2020\|EPI_ISL_519717\|2020 | ···················· | ···················· | ···················· | ···················· | ···················· | ···················· | |
| G | 6949 | hCoV-19/Australia/VIC4422/2020\|EPI_ISL_519718\|2020 | ···················· | ···················· | ···················· | ···················· | ···················· | ···················· | |
| G | 6950 | hCoV-19/Australia/VIC4423/2020\|EPI_ISL_519719\|2020 | ···················· | ···················· | ···················· | ···················· | ···················· | ···················· | |
| G | 6951 | hCoV-19/Australia/VIC4424/2020\|EPI_ISL_519720\|2020 | ···················· | ···················· | ···················· | ···················· | ···················· | ···················· | |
| G | 6952 | hCoV-19/Australia/VIC4425/2020\|EPI_ISL_519721\|2020 | ···················· | ···················· | ···················· | ···················· | ···················· | ···················· | |
| G | 6953 | hCoV-19/Australia/VIC4426/2020\|EPI_ISL_519722\|2020 | ···················· | ···················· | ···················· | ···················· | ···················· | ···················· | |
| G | 6954 | hCoV-19/Australia/VIC4427/2020\|EPI_ISL_519723\|2020 | ···················· | ···················· | ···················· | ···················· | ···················· | ···················· | |
| G | 6955 | hCoV-19/Australia/VIC4428/2020\|EPI_ISL_519724\|2020 | ···················· | ···················· | ···················· | ···················· | ···················· | ···················· | |
| G | 6956 | hCoV-19/Australia/VIC4429/2020\|EPI_ISL_519725\|2020 | ···················· | ···················· | ···················· | ···················· | ···················· | ···················· | |
| G | 6957 | hCoV-19/Australia/VIC4430/2020\|EPI_ISL_519726\|2020 | ···················· | ···················· | ···················· | ···················· | ···················· | ···················· | |
| G | 6958 | hCoV-19/Australia/VIC4431/2020\|EPI_ISL_519727\|2020 | ···················· | ···················· | ···················· | ···················· | ···················· | ···················· | |
| G | 6959 | hCoV-19/Australia/VIC4432/2020\|EPI_ISL_519728\|2020 | ···················· | ···················· | ···················· | ···················· | ···················· | ···················· | |
| G | 6960 | hCoV-19/Australia/VIC4433/2020\|EPI_ISL_519729\|2020 | ···················· | ···················· | ···················· | ···················· | ···················· | ···················· | |
| G | 6961 | hCoV-19/Australia/VIC4434/2020\|EPI_ISL_519730\|2020 | ···················· | ···················· | ···················· | ···················· | ···················· | ···················· | |
| G | 6962 | hCoV-19/Australia/VIC4435/2020\|EPI_ISL_519731\|2020 | ···················· | ···················· | ···················· | ···················· | ···················· | ···················· | |
| G | 6963 | hCoV-19/Australia/VIC4436/2020\|EPI_ISL_519732\|2020 | ···················· | ···················· | ···················· | ···················· | ···················· | ···················· | |
| G | 6964 | hCoV-19/Australia/VIC4437/2020\|EPI_ISL_519733\|2020 | ···················· | ···················· | ···················· | ···················· | ···················· | ···················· | |
| G | 6965 | hCoV-19/Australia/VIC4438/2020\|EPI_ISL_519734\|2020 | ···················· | ···················· | ···················· | ···················· | ···················· | ···················· | |
| G | 6966 | hCoV-19/Australia/VIC4439/2020\|EPI_ISL_519735\|2020 | ···················· | ···················· | ···················· | ···················· | ···················· | ···················· | |
| G | 6967 | hCoV-19/Australia/VIC4440/2020\|EPI_ISL_519736\|2020 | ···················· | ···················· | ···················· | ···················· | ···················· | ···················· | |
| G | 6968 | hCoV-19/Australia/VIC4441/2020\|EPI_ISL_519737\|2020 | ···················· | ···················· | ···················· | ···················· | ···················· | ···················· | |
| G | 6969 | hCoV-19/Australia/VIC4442/2020\|EPI_ISL_519738\|2020 | ···················· | ···················· | ···················· | ···················· | ···················· | ···················· | |
| G | 6970 | hCoV-19/Australia/VIC4443/2020\|EPI_ISL_519739\|2020 | ···················· | ···················· | ···················· | ···················· | ···················· | ···················· | |
| G | 6971 | hCoV-19/United Arab Emirates/L3290259618/2020\|EPI_ | ···················· | ···················· | ···················· | ···················· | ···················· | ···················· | |
| G | 6972 | hCoV-19/United Arab Emirates/L3290262866/2020\|EPI_ | ···················· | ···················· | ···················· | ···················· | ···················· | ···················· | |
| G | 6973 | hCoV-19/United Arab Emirates/L3290263078/2020\|EPI_ | ···················· | ···················· | ···················· | ···················· | ···················· | ···················· | |
| G | 6974 | hCoV-19/United Arab Emirates/L3290263088/2020\|EPI_ | ···················· | ···················· | ···················· | ···················· | ···················· | ···················· | |
| G | 6975 | hCoV-19/United Arab Emirates/L3290264100/2020\|EPI_ | ···················· | ···················· | ···················· | ···················· | ···················· | ···················· | |
| G | 6976 | hCoV-19/United Arab Emirates/L3290294240/2020\|EPI_ | ···················· | ···················· | ···················· | ···················· | ···················· | ···················· | |
| G | 6977 | hCoV-19/United Arab Emirates/L3290314014/2020\|EPI_ | ···················· | ···················· | ···················· | ···················· | ···················· | ···················· | |
| G | 6978 | hCoV-19/United Arab Emirates/L3290324509/2020\|EPI_ | ···················· | ···················· | ···················· | ···················· | ···················· | ···················· | |
| G | 6979 | hCoV-19/United Arab Emirates/L3290361549/2020\|EPI_ | ···················· | ···················· | ···················· | ···················· | ···················· | ···················· | |
| G | 6980 | hCoV-19/United Arab Emirates/L3290362429/2020\|EPI_ | ···················· | ···················· | ···················· | ···················· | ···················· | ···················· | |
| G | 6981 | hCoV-19/United Arab Emirates/L3290363526/2020\|EPI_ | ···················· | ···················· | ···················· | ···················· | ···················· | ···················· | |
| G | 6982 | hCoV-19/United Arab Emirates/L3300054450/2020\|EPI_ | ···················· | ···················· | ···················· | ···················· | ···················· | ···················· | |
| G | 6983 | hCoV-19/United Arab Emirates/L3300059655/2020\|EPI_ | ···················· | ···················· | ···················· | ···················· | ···················· | ···················· | |
| G | 6984 | hCoV-19/United Arab Emirates/L3300061195/2020\|EPI_ | ···················· | ···················· | ···················· | ···················· | ···················· | ···················· | |
| G | 6985 | hCoV-19/United Arab Emirates/L3300061831/2020\|EPI_ | ···················· | ···················· | ···················· | ···················· | ···················· | ···················· | |
| G | 6986 | hCoV-19/United Arab Emirates/L3300068662/2020\|EPI_ | ···················· | ···················· | ···················· | ···················· | ···················· | ···················· | |
| G | 6987 | hCoV-19/United Arab Emirates/L46302981/2020\|EPI_IS | ···················· | ···················· | ···················· | ···················· | ···················· | ···················· | |
| G | 6988 | hCoV-19/United Arab Emirates/L920066831/2020\|EPI_I | ···················· | ···················· | ···················· | ···················· | ···················· | ···················· | |
| G | 6989 | hCoV-19/United Arab Emirates/L920331360/2020\|EPI_I | ···················· | ···················· | ···················· | ···················· | ···················· | ···················· | |
| G | 6990 | hCoV-19/United Arab Emirates/L920626247/2020\|EPI_I | ···················· | ···················· | ···················· | ···················· | ···················· | ···················· | |
| G | 6991 | hCoV-19/Australia/VIC3090/2020\|EPI_ISL_520797\|2020 | ···················· | ···················· | ···················· | ···················· | ···················· | ···················· | |
| G | 6992 | hCoV-19/Australia/VIC3091/2020\|EPI_ISL_520798\|2020 | ···················· | ···················· | ···················· | ···················· | ···················· | ···················· | |
| G | 6993 | hCoV-19/Australia/VIC3092/2020\|EPI_ISL_520799\|2020 | ···················· | ···················· | ···················· | ···················· | NNN················· | ···················· | 1 |
| G | 6994 | hCoV-19/USA/UT-08422/2020\|EPI_ISL_524200\|2020-05-2 | ···················· | ···················· | ···················· | ···················· | ···················· | ···················· | |
| G | 6995 | hCoV-19/USA/UT-08425/2020\|EPI_ISL_524201\|2020-05-1 | ···················· | ···················· | ···················· | ···················· | ···················· | ···················· | |
| G | 6996 | hCoV-19/USA/UT-08426/2020\|EPI_ISL_524202\|2020-05-1 | ···················· | ···················· | ···················· | ···················· | ···················· | ···················· | |
| G | 6997 | hCoV-19/USA/UT-08427/2020\|EPI_ISL_524203\|2020-05-1 | ···················· | ···················· | ···················· | ···················· | ···················· | ···················· | |
| G | 6998 | hCoV-19/USA/UT-08428/2020\|EPI_ISL_524204\|2020-05-2 | ···················· | ···················· | ···················· | ···················· | ···················· | ···················· | |
| G | 6999 | hCoV-19/USA/UT-08430/2020\|EPI_ISL_524206\|2020-05-2 | ···················· | ···················· | ···················· | ···················· | ···················· | ···················· | |
| G | 7000 | hCoV-19/USA/UT-08481/2020\|EPI_ISL_524250\|2020-05-2 | ···················· | ···················· | ···················· | ···················· | ·········A·········· | ···················· | |
| G | 7001 | hCoV-19/USA/UT-08482/2020\|EPI_ISL_524251\|2020-05-1 | ···················· | ···················· | ···················· | ···················· | ···················· | ···················· | |
| G | 7002 | hCoV-19/USA/UT-08484/2020\|EPI_ISL_524253\|2020-05-1 | ···················· | ···················· | ···················· | ···················· | ···················· | ···················· | |
| G | 7003 | hCoV-19/USA/UT-08486/2020\|EPI_ISL_524255\|2020-05-1 | ···················· | ···················· | ···················· | ···················· | ···················· | ···················· | |
| G | 7004 | hCoV-19/USA/UT-08487/2020\|EPI_ISL_524256\|2020-05-2 | ···················· | ···················· | ···················· | ···················· | ···················· | ···················· | |
| G | 7005 | hCoV-19/USA/UT-08489/2020\|EPI_ISL_524257\|2020-05-2 | ···················· | ···················· | ···················· | ···················· | ···················· | ···················· | |
| G | 7006 | hCoV-19/USA/UT-08492/2020\|EPI_ISL_524260\|2020-05-2 | ···················· | ···················· | ···················· | ···················· | ···················· | ···················· | |
| G | 7007 | hCoV-19/USA/UT-08494/2020\|EPI_ISL_524261\|2020-05-2 | ···················· | ···················· | ···················· | ···················· | ···················· | ···················· | |
| G | 7008 | hCoV-19/USA/UT-08495/2020\|EPI_ISL_524262\|2020-05-1 | ···················· | ···················· | ···················· | ···················· | ···················· | ···················· | |
| G | 7009 | hCoV-19/USA/UT-08496/2020\|EPI_ISL_524263\|2020-05-1 | ···················· | ···················· | ···················· | ···················· | ···················· | ···················· | |
| G | 7010 | hCoV-19/USA/UT-08498/2020\|EPI_ISL_524265\|2020-05-1 | ···················· | ···················· | ···················· | ···················· | ···················· | ···················· | |
| G | 7011 | hCoV-19/USA/UT-08499/2020\|EPI_ISL_524266\|2020-05-1 | ···················· | ···················· | ···················· | ···················· | ···················· | ···················· | |
| G | 7012 | hCoV-19/USA/UT-08503/2020\|EPI_ISL_524269\|2020-05-1 | ···················· | ···················· | ···················· | ···················· | ···················· | ···················· | |
| G | 7013 | hCoV-19/USA/UT-08506/2020\|EPI_ISL_524271\|2020-05-1 | ···················· | ···················· | ···················· | ···················· | ···················· | ···················· | |
| G | 7014 | hCoV-19/USA/UT-08507/2020\|EPI_ISL_524272\|2020-05-1 | ···················· | ···················· | ···················· | ···················· | ···················· | ···················· | |
| G | 7015 | hCoV-19/USA/UT-08510/2020\|EPI_ISL_524274\|2020-05-1 | ···················· | ···················· | ···················· | ···················· | ···C················ | ···················· | |
| G | 7016 | hCoV-19/USA/UT-08511/2020\|EPI_ISL_524275\|2020-05-1 | ···················· | ···················· | ···················· | ···················· | ···················· | ···················· | |
| G | 7017 | hCoV-19/USA/UT-08514/2020\|EPI_ISL_524277\|2020-05-1 | ···················· | ···················· | ···················· | ···················· | ···················· | ···················· | |
| G | 7018 | hCoV-19/USA/UT-08516/2020\|EPI_ISL_524279\|2020-05-2 | ···················· | ···················· | ···················· | ···················· | ···················· | ···················· | |
| G | 7019 | hCoV-19/USA/UT-08517/2020\|EPI_ISL_524280\|2020-05-1 | ···················· | ···················· | ···················· | ···················· | ···················· | ···················· | |
| G | 7020 | hCoV-19/USA/UT-08518/2020\|EPI_ISL_524281\|2020-05-1 | ···················· | ···················· | ···················· | ···················· | ···················· | ···················· | |
| G | 7021 | hCoV-19/USA/UT-08520/2020\|EPI_ISL_524283\|2020-05-1 | ···················· | ···················· | ···················· | ···················· | ···················· | ···················· | |
| G | 7022 | hCoV-19/USA/UT-08521/2020\|EPI_ISL_524284\|2020-05-1 | ···················· | ···················· | ···················· | ···················· | ···················· | ···················· | |
| G | 7023 | hCoV-19/USA/UT-08522/2020\|EPI_ISL_524285\|2020-05-1 | ···················· | ···················· | ···················· | ···················· | ···················· | ···················· | |

| | | | | | | | |
|---|---|---|---|---|---|---|---|
| G | 7024 hCoV-19/USA/UT-08525/2020\|EPI_ISL_524288\|2020-05-1 | ···················· | ···················· | ····················· | ···················· | ···················· | ···················· |
| G | 7025 hCoV-19/USA/UT-08526/2020\|EPI_ISL_524289\|2020-05-2 | ···················· | ···················· | ····················· | ···················· | ···················· | ···················· |
| G | 7026 hCoV-19/USA/UT-08527/2020\|EPI_ISL_524290\|2020-05-2 | ···················· | ·A·················· | ····················· | ···················· | ···················· | ···················· |
| G | 7027 hCoV-19/USA/UT-08528/2020\|EPI_ISL_524291\|2020-05-1 | ···················· | ···················· | ····················· | ···················· | ···················· | ···················· |
| G | 7028 hCoV-19/USA/UT-08529/2020\|EPI_ISL_524292\|2020-05-1 | ···················· | ···················· | ····················· | ···················· | ···················· | ···················· |
| G | 7029 hCoV-19/USA/UT-08530/2020\|EPI_ISL_524293\|2020-05-1 | ···················· | ···················· | ····················· | ···················· | ···················· | ···················· |
| G | 7030 hCoV-19/USA/UT-08534/2020\|EPI_ISL_524295\|2020-05-1 | ···················· | ···················· | ····················· | ···················· | ···················· | ···················· |
| G | 7031 hCoV-19/USA/UT-08535/2020\|EPI_ISL_524296\|2020-05-1 | ···················· | ···················· | ····················· | ···················· | ···················· | ···················· |
| G | 7032 hCoV-19/USA/UT-08536/2020\|EPI_ISL_524297\|2020-05-1 | ···················· | ···················· | ····················· | ···················· | ···················· | ···················· |
| G | 7033 hCoV-19/USA/UT-08538/2020\|EPI_ISL_524299\|2020-05-1 | ···················· | ···················· | ····················· | ···················· | ···················· | ···················· |
| G | 7034 hCoV-19/France/ARA-06391/2020\|EPI_ISL_525536\|2020- | ···················· | ···················· | ····················· | ···················· | ···················· | ···················· |
| G | 7035 hCoV-19/France/ARA-10194/2020\|EPI_ISL_525537\|2020- | ···················· | ···················· | ····················· | ···················· | ···················· | ···················· |
| G | 7036 hCoV-19/USA/NY-Wadsworth-11244-01/2020\|EPI_ISL_525 | ···················· | ···················· | ····················· | ···················· | ···················· | ···················· |
| G | 7037 hCoV-19/USA/NY-Wadsworth-11513-01/2020\|EPI_ISL_525 | ···················· | ···················· | ····················· | ···················· | ···················· | ···················· |
| G | 7038 hCoV-19/USA/NY-Wadsworth-11537-01/2020\|EPI_ISL_525 | ···················· | ···················· | ····················· | ···················· | ···················· | ···················· |
| G | 7039 hCoV-19/USA/NY-Wadsworth-11538-01/2020\|EPI_ISL_525 | ···················· | ···················· | ····················· | ···················· | ···················· | ···················· |
| G | 7040 hCoV-19/USA/NY-Wadsworth-11612-01/2020\|EPI_ISL_525 | ···················· | ···················· | ····················· | ···················· | ···················· | ···················· |
| G | 7041 hCoV-19/USA/NY-Wadsworth-11681-01/2020\|EPI_ISL_525 | ···················· | ···················· | ····················· | ···················· | ···················· | ···················· |
| G | 7042 hCoV-19/USA/NY-Wadsworth-11690-01/2020\|EPI_ISL_525 | ···················· | ···················· | ····················· | ···················· | ···················· | ···················· |
| G | 7043 hCoV-19/USA/NY-Wadsworth-11719-01/2020\|EPI_ISL_525 | ···················· | ···················· | ····················· | ···················· | ···················· | ···················· |
| G | 7044 hCoV-19/USA/NY-Wadsworth-11789-01/2020\|EPI_ISL_525 | ···················· | ···················· | ····················· | ···················· | ···················· | ···················· |
| G | 7045 hCoV-19/USA/NY-Wadsworth-12211-01/2020\|EPI_ISL_525 | ···················· | ···················· | ····················· | ···················· | ···················· | ···················· |
| G | 7046 hCoV-19/USA/NY-Wadsworth-12669-01/2020\|EPI_ISL_525 | ···················· | ···················· | ····················· | ···················· | ···················· | ···················· |
| G | 7047 hCoV-19/USA/NY-Wadsworth-12692-01/2020\|EPI_ISL_525 | ···················· | ···················· | ····················· | ···················· | ···················· | ···················· |
| G | 7048 hCoV-19/USA/NY-Wadsworth-12767-01/2020\|EPI_ISL_525 | ···················· | ···················· | ····················· | ···················· | ···················· | ···················· |
| G | 7049 hCoV-19/USA/NY-Wadsworth-12806-01/2020\|EPI_ISL_525 | ···················· | ···················· | ····················· | ···················· | ···················· | ···················· |
| G | 7050 hCoV-19/USA/NY-Wadsworth-12813-01/2020\|EPI_ISL_525 | ···················· | ···················· | ····················· | ···················· | ···················· | ···················· |
| G | 7051 hCoV-19/USA/NY-Wadsworth-12824-01/2020\|EPI_ISL_525 | ···················· | ···················· | ····················· | ···················· | ···················· | ···················· |
| G | 7052 hCoV-19/USA/NY-Wadsworth-12848-01/2020\|EPI_ISL_525 | ···················· | ···················· | ····················· | ···················· | ···················· | ···················· |
| G | 7053 hCoV-19/USA/NY-Wadsworth-13245-01/2020\|EPI_ISL_525 | ···················· | ···················· | ····················· | ···················· | ···················· | ···················· |
| G | 7054 hCoV-19/USA/VA-DCLS-0608/2020\|EPI_ISL_526825\|2020- | ···················· | ···················· | ····················· | ···················· | ···················· | ···················· |
| G | 7055 hCoV-19/USA/VA-DCLS-0609/2020\|EPI_ISL_526826\|2020- | ···················· | ···················· | ····················· | ···················· | ···················· | ···················· |
| G | 7056 hCoV-19/USA/VA-DCLS-0610/2020\|EPI_ISL_526827\|2020- | ···················· | ···················· | ····················· | ···················· | ···················· | ···················· |
| G | 7057 hCoV-19/USA/VA-DCLS-0612/2020\|EPI_ISL_526828\|2020- | ···················· | ···················· | ····················· | ···················· | ···················· | ···················· |
| G | 7058 hCoV-19/USA/VA-DCLS-0613/2020\|EPI_ISL_526829\|2020- | ···················· | ···················· | ····················· | ···················· | ···················· | ···················· |
| G | 7059 hCoV-19/USA/VA-DCLS-0614/2020\|EPI_ISL_526830\|2020- | ···················· | ···················· | ····················· | ···················· | ···················· | ···················· |
| G | 7060 hCoV-19/USA/VA-DCLS-0616/2020\|EPI_ISL_526831\|2020- | ···················· | ···················· | ····················· | ···················· | ···················· | ···················· |
| G | 7061 hCoV-19/USA/VA-DCLS-0617/2020\|EPI_ISL_526832\|2020- | ···················· | ···················· | ····················· | ···················· | ···················· | ···················· |
| G | 7062 hCoV-19/USA/VA-DCLS-0619/2020\|EPI_ISL_526833\|2020- | ···················· | ···················· | ····················· | ···················· | ···················· | ···················· |
| G | 7063 hCoV-19/USA/VA-DCLS-0621/2020\|EPI_ISL_526834\|2020- | ···················· | ···················· | ····················· | ···················· | ···················· | ···················· |
| G | 7064 hCoV-19/USA/VA-DCLS-0622/2020\|EPI_ISL_526835\|2020- | ···················· | ···················· | ····················· | ···················· | ···················· | ···················· |
| G | 7065 hCoV-19/USA/VA-DCLS-0659/2020\|EPI_ISL_526836\|2020- | ···················· | ···················· | ····················· | ···················· | ···················· | ···················· |
| G | 7066 hCoV-19/USA/VA-DCLS-0660/2020\|EPI_ISL_526837\|2020- | ···················· | ···················· | ····················· | ···················· | ···················· | ···················· |
| G | 7067 hCoV-19/USA/VA-DCLS-0661/2020\|EPI_ISL_526838\|2020- | ···················· | ···················· | ····················· | ···················· | ···················· | ···················· |
| G | 7068 hCoV-19/USA/VA-DCLS-0662/2020\|EPI_ISL_526839\|2020- | ···················· | ···················· | ····················· | ···················· | ···················· | ···················· |
| G | 7069 hCoV-19/USA/VA-DCLS-0664/2020\|EPI_ISL_526840\|2020- | ···················· | ···················· | ····················· | ···················· | ···················· | ···················· |
| G | 7070 hCoV-19/USA/VA-DCLS-0665/2020\|EPI_ISL_526841\|2020- | ···················· | ···················· | ····················· | ···················· | ···················· | ···················· |
| G | 7071 hCoV-19/USA/VA-DCLS-0666/2020\|EPI_ISL_526842\|2020- | ···················· | ···················· | ····················· | ···················· | ···················· | ···················· |
| G | 7072 hCoV-19/USA/VA-DCLS-0667/2020\|EPI_ISL_526843\|2020- | ···················· | ···················· | ····················· | ···················· | ···················· | ···················· |
| G | 7073 hCoV-19/USA/VA-DCLS-0721/2020\|EPI_ISL_526844\|2020- | ···················· | ···················· | ····················· | ···················· | ···················· | ···················· |
| G | 7074 hCoV-19/USA/VA-DCLS-0722/2020\|EPI_ISL_526845\|2020- | ···········T········ | ···················· | ····················· | ···················· | ···················· | ···················· |
| G | 7075 hCoV-19/USA/VA-DCLS-0723/2020\|EPI_ISL_526846\|2020- | ···········T········ | ···················· | ····················· | ···················· | ···················· | ···················· |
| G | 7076 hCoV-19/USA/VA-DCLS-0724/2020\|EPI_ISL_526847\|2020- | ···········T········ | ···················· | ····················· | ···················· | ···················· | ···················· |
| G | 7077 hCoV-19/USA/VA-DCLS-0725/2020\|EPI_ISL_526848\|2020- | ···········T········ | ···················· | ····················· | ···················· | ···················· | ···················· |
| G | 7078 hCoV-19/USA/VA-DCLS-0726/2020\|EPI_ISL_526849\|2020- | ···········T········ | ···················· | ····················· | ···················· | ···················· | ···················· |
| G | 7079 hCoV-19/USA/VA-DCLS-0727/2020\|EPI_ISL_526850\|2020- | ···········T········ | ···················· | ····················· | ···················· | ···················· | ···················· |
| G | 7080 hCoV-19/USA/VA-DCLS-0728/2020\|EPI_ISL_526851\|2020- | ···········T········ | ···················· | ····················· | ···················· | ···················· | ···················· |
| G | 7081 hCoV-19/USA/VA-DCLS-0729/2020\|EPI_ISL_526852\|2020- | ···········T········ | ···················· | ····················· | ···················· | ···················· | ···················· |
| G | 7082 hCoV-19/USA/VA-DCLS-0731/2020\|EPI_ISL_526853\|2020- | ···········T········ | ···················· | ····················· | ···················· | ···················· | ···················· |
| G | 7083 hCoV-19/USA/VA-DCLS-0732/2020\|EPI_ISL_526854\|2020- | ···········T········ | ···················· | ····················· | ···················· | ···················· | ···················· |
| G | 7084 hCoV-19/USA/VA-DCLS-0733/2020\|EPI_ISL_526855\|2020- | ···········T········ | ···················· | ····················· | ···················· | ···················· | ···················· |
| G | 7085 hCoV-19/USA/VA-DCLS-0734/2020\|EPI_ISL_526856\|2020- | ···········T········ | ···················· | ····················· | ···················· | ···················· | ···················· |
| G | 7086 hCoV-19/USA/VA-DCLS-0736/2020\|EPI_ISL_526857\|2020- | ···················· | ·A·················· | ····················· | ···················· | ···················· | ···················· |
| G | 7087 hCoV-19/USA/VA-DCLS-0737/2020\|EPI_ISL_526858\|2020- | ···················· | ·A·················· | ····················· | ···················· | ···················· | ···················· |
| G | 7088 hCoV-19/USA/VA-DCLS-0738/2020\|EPI_ISL_526859\|2020- | ···················· | ·A·················· | ····················· | ···················· | ···················· | ···················· |
| G | 7089 hCoV-19/USA/VA-DCLS-0739/2020\|EPI_ISL_526860\|2020- | ···················· | ·A·················· | ····················· | ···················· | ···················· | ···················· |
| G | 7090 hCoV-19/USA/VA-DCLS-0740/2020\|EPI_ISL_526861\|2020- | ···················· | ·A·················· | ····················· | ···················· | ···················· | ···················· |
| G | 7091 hCoV-19/USA/VA-DCLS-0741/2020\|EPI_ISL_526862\|2020- | ···················· | ·A·················· | ····················· | ···················· | ···················· | ···················· |
| G | 7092 hCoV-19/USA/VA-DCLS-0743/2020\|EPI_ISL_526863\|2020- | ···················· | ···················· | ····················· | ···················· | ···················· | ···················· |
| G | 7093 hCoV-19/USA/VA-DCLS-0746/2020\|EPI_ISL_526864\|2020- | ···················· | ···················· | ····················· | ···················· | ···················· | ···················· |
| G | 7094 hCoV-19/USA/VA-DCLS-0748/2020\|EPI_ISL_526865\|2020- | ···················· | ···················· | ····················· | ···················· | ···················· | ···················· |
| G | 7095 hCoV-19/USA/VA-DCLS-0749/2020\|EPI_ISL_526866\|2020- | ···················· | ···················· | ····················· | ···················· | ···················· | ···················· |
| G | 7096 hCoV-19/USA/VA-DCLS-0750/2020\|EPI_ISL_526867\|2020- | ···················· | ···················· | ····················· | ···················· | ···················· | ···················· |
| G | 7097 hCoV-19/USA/VA-DCLS-0752/2020\|EPI_ISL_526868\|2020- | ···················· | ···················· | ····················· | ···················· | ···················· | ···················· |
| G | 7098 hCoV-19/Japan/QH-329-037/2020\|EPI_ISL_529135\|2020- | ···················· | ···················· | ····················· | ···················· | ···················· | ···················· |
| G | 7099 hCoV-19/Indonesia/EJ-ITD-8402NT/2020\|EPI_ISL_52913 | ···················· | ···················· | ····················· | ···················· | ···················· | ···················· |
| G | 7100 hCoV-19/Spain/MD-HLP-112/2020\|EPI_ISL_530041\|2020- | ···················· | ···················· | ····················· | ···················· | ···················· | ···················· |
| G | 7101 hCoV-19/Spain/MD-HLP-113/2020\|EPI_ISL_530042\|2020- | ···················· | ···················· | ····················· | ···················· | ···················· | ···················· |
| G | 7102 hCoV-19/Spain/MD-HLP-114/2020\|EPI_ISL_530043\|2020- | ···················· | ···················· | ····················· | ···················· | ···················· | ···················· |
| G | 7103 hCoV-19/Spain/MD-HLP-40/2020\|EPI_ISL_530044\|2020-0 | ···················· | ···················· | ····················· | ···················· | ···················· | ···················· |

| | | | | | | | | |
|---|---|---|---|---|---|---|---|---|
| G | 7104 | hCoV-19/Spain/MD-HLP-41/2020\|EPI_ISL_530045\|2020-0 | ···················· | ···················· | ····················· | ···················· | ···················· | ···················· |
| G | 7105 | hCoV-19/Spain/MD-HLP-42/2020\|EPI_ISL_530046\|2020-0 | ···················· | ···················· | ····················· | ···················· | ···················· | ···················· |
| G | 7106 | hCoV-19/Spain/MD-HLP-43/2020\|EPI_ISL_530047\|2020-0 | ···················· | ···················· | ····················· | ···················· | ···················· | ···················· |
| G | 7107 | hCoV-19/Spain/MD-HLP-44/2020\|EPI_ISL_530048\|2020-0 | ···················· | ···················· | ····················· | ···················· | ···················· | ···················· |
| G | 7108 | hCoV-19/Spain/MD-HLP-45/2020\|EPI_ISL_530049\|2020-0 | ···················· | ···················· | ····················· | ···················· | ···················· | ···················· |
| G | 7109 | hCoV-19/Spain/MD-HLP-46/2020\|EPI_ISL_530050\|2020-0 | ···················· | ···················· | ····················· | ···················· | ···················· | ···················· |
| G | 7110 | hCoV-19/Spain/MD-HLP-48/2020\|EPI_ISL_530051\|2020-0 | ···················· | ···················· | ····················· | ···················· | ···················· | ···················· |
| G | 7111 | hCoV-19/Spain/MD-HLP-49/2020\|EPI_ISL_530052\|2020-0 | ···················· | ···················· | ····················· | ···················· | ···················· | ···················· |
| G | 7112 | hCoV-19/Spain/MD-HLP-50/2020\|EPI_ISL_530053\|2020-0 | ···················· | ···················· | ···········T········· | ···················· | ···················· | ···················· |
| G | 7113 | hCoV-19/Spain/MD-HLP-51/2020\|EPI_ISL_530054\|2020-0 | ···················· | ···················· | ····················· | ···················· | ···················· | ···················· |
| G | 7114 | hCoV-19/Spain/MD-HLP-52/2020\|EPI_ISL_530055\|2020-0 | ···················· | ···················· | ····················· | ···················· | ···················· | ···················· |
| G | 7115 | hCoV-19/Spain/MD-HLP-53/2020\|EPI_ISL_530056\|2020-0 | ···················· | ···················· | ····················· | ···················· | ···················· | ···················· |
| G | 7116 | hCoV-19/Spain/MD-HLP-54/2020\|EPI_ISL_530057\|2020-0 | ···················· | ···················· | ····················· | ···················· | ···················· | ···················· |
| G | 7117 | hCoV-19/Spain/MD-HLP-55/2020\|EPI_ISL_530058\|2020-0 | ···················· | ···················· | ····················· | ···················· | ···················· | ···················· |
| G | 7118 | hCoV-19/Spain/MD-HLP-56/2020\|EPI_ISL_530059\|2020-0 | ···················· | ···················· | ····················· | ···················· | ···················· | ···················· |
| G | 7119 | hCoV-19/Spain/MD-HLP-57/2020\|EPI_ISL_530060\|2020-0 | ···················· | ···················· | ····················· | ···················· | ···················· | ···················· |
| G | 7120 | hCoV-19/Spain/MD-HLP-58/2020\|EPI_ISL_530061\|2020-0 | ···················· | ···················· | ····················· | ···················· | ···················· | ···················· |
| G | 7121 | hCoV-19/Spain/MD-HLP-59/2020\|EPI_ISL_530062\|2020-0 | ···················· | ···················· | ····················· | ···················· | ···················· | ···················· |
| G | 7122 | hCoV-19/Spain/MD-HLP-60/2020\|EPI_ISL_530063\|2020-0 | ···················· | ···················· | ····················· | ···················· | ···················· | ···················· |
| G | 7123 | hCoV-19/Spain/MD-HLP-61/2020\|EPI_ISL_530064\|2020-0 | ···················· | ···················· | ····················· | ···················· | ···················· | ···················· |
| G | 7124 | hCoV-19/Spain/MD-HLP-62/2020\|EPI_ISL_530065\|2020-0 | ···················· | ···················· | ····················· | ···················· | ···················· | ···················· |
| G | 7125 | hCoV-19/Spain/MD-HLP-63/2020\|EPI_ISL_530066\|2020-0 | ···················· | ···················· | ····················· | ···················· | ···················· | ···················· |
| G | 7126 | hCoV-19/Spain/MD-HLP-64/2020\|EPI_ISL_530067\|2020-0 | ···················· | ···················· | ····················· | ···················· | ···················· | ···················· |
| G | 7127 | hCoV-19/Spain/MD-HLP-65/2020\|EPI_ISL_530068\|2020-0 | ···················· | ···················· | ····················· | ···················· | ···················· | ···················· |
| G | 7128 | hCoV-19/Spain/MD-HLP-68/2020\|EPI_ISL_530069\|2020-0 | ···················· | ···················· | ····················· | ···················· | ···················· | ···················· |
| G | 7129 | hCoV-19/Spain/MD-HLP-71/2020\|EPI_ISL_530070\|2020-0 | ···················· | ···················· | ····················· | ···················· | ···················· | ···················· |
| G | 7130 | hCoV-19/Spain/MD-HLP-72/2020\|EPI_ISL_530071\|2020-0 | ···················· | ···················· | ····················· | ···················· | ···················· | ···················· |
| G | 7131 | hCoV-19/Spain/MD-HLP-73/2020\|EPI_ISL_530072\|2020-0 | ···················· | ···················· | ····················· | ···················· | ···················· | ···················· |
| G | 7132 | hCoV-19/Spain/MD-HLP-74/2020\|EPI_ISL_530073\|2020-0 | ···················· | ···················· | ····················· | ···················· | ···················· | ···················· |
| G | 7133 | hCoV-19/Spain/MD-HLP-77/2020\|EPI_ISL_530074\|2020-0 | ···················· | ···················· | ····················· | ···················· | ···················· | ···················· |
| G | 7134 | hCoV-19/Spain/MD-HLP-78/2020\|EPI_ISL_530075\|2020-0 | ···················· | ···················· | ····················· | ···················· | ···················· | ···················· |
| G | 7135 | hCoV-19/Spain/MD-HLP-80/2020\|EPI_ISL_530077\|2020-0 | ···················· | ···················· | ····················· | ···················· | ···················· | ···················· |
| G | 7136 | hCoV-19/Spain/MD-HLP-81/2020\|EPI_ISL_530078\|2020-0 | ···················· | ···················· | ····················· | ···················· | ···················· | ···················· |
| G | 7137 | hCoV-19/Spain/MD-HLP-R82/2020\|EPI_ISL_530079\|2020- | ···················· | ···················· | ····················· | ···················· | ···················· | ···················· |
| G | 7138 | hCoV-19/Spain/MD-HLP-83/2020\|EPI_ISL_530080\|2020-0 | ···················· | ···················· | ····················· | ···················· | ···················· | ···················· |
| G | 7139 | hCoV-19/Spain/MD-HLP-84/2020\|EPI_ISL_530081\|2020-0 | ···················· | ···················· | ····················· | ···················· | ···················· | ···················· |
| G | 7140 | hCoV-19/Spain/MD-HLP-85/2020\|EPI_ISL_530082\|2020-0 | ···················· | ···················· | ····················· | ···················· | ···················· | ···················· |
| G | 7141 | hCoV-19/Spain/MD-HLP-86/2020\|EPI_ISL_530083\|2020-0 | ···················· | ···················· | ····················· | ···················· | ···················· | ···················· |
| G | 7142 | hCoV-19/Spain/MD-HLP-87/2020\|EPI_ISL_530084\|2020-0 | ···················· | ···················· | ····················· | ···················· | ···················· | ···················· |
| G | 7143 | hCoV-19/Spain/MD-HLP-88/2020\|EPI_ISL_530085\|2020-0 | ···················· | ···················· | ····················· | ···················· | ···················· | ···················· |
| G | 7144 | hCoV-19/Spain/MD-HLP-89/2020\|EPI_ISL_530086\|2020-0 | ···················· | ···················· | ····················· | ···················· | ···················· | ···················· |
| G | 7145 | hCoV-19/Spain/MD-HLP-90/2020\|EPI_ISL_530087\|2020-0 | ···················· | ···················· | ····················· | ···················· | ···················· | ···················· |
| G | 7146 | hCoV-19/Spain/MD-HLP-92/2020\|EPI_ISL_530088\|2020-0 | ···················· | ···················· | ····················· | ···················· | ···················· | ···················· |
| G | 7147 | hCoV-19/Spain/MD-HLP-93/2020\|EPI_ISL_530089\|2020-0 | ···················· | ···················· | ····················· | ···················· | ···················· | ···················· |
| G | 7148 | hCoV-19/Spain/MD-HLP-R50/2020\|EPI_ISL_530100\|2020- | ···················· | ···A················ | ····················· | ···················· | ···················· | ···················· |
| G | 7149 | hCoV-19/Spain/MD-HLP-R51/2020\|EPI_ISL_530101\|2020- | ···················· | ···················· | ····················· | ···················· | ···················· | ···················· |
| G | 7150 | hCoV-19/Spain/MD-HLP-R52/2020\|EPI_ISL_530102\|2020- | ···················· | ···················· | ····················· | ···················· | ···················· | ···················· |
| G | 7151 | hCoV-19/Spain/MD-HLP-R53/2020\|EPI_ISL_530103\|2020- | ···················· | ···················· | ····················· | ···················· | ···················· | ···················· |
| G | 7152 | hCoV-19/Spain/MD-HLP-R56/2020\|EPI_ISL_530104\|2020- | ···················· | ···················· | ····················· | ···················· | ···················· | ···················· |
| G | 7153 | hCoV-19/Spain/MD-HLP-R59/2020\|EPI_ISL_530105\|2020- | ···················· | ···················· | ····················· | ···················· | ···················· | ···················· |
| G | 7154 | hCoV-19/Spain/MD-HLP-R60/2020\|EPI_ISL_530106\|2020- | ···················· | ···················· | ····················· | ···················· | ···················· | ···················· |
| G | 7155 | hCoV-19/Spain/MD-HLP-R61/2020\|EPI_ISL_530107\|2020- | ···················· | ···················· | ····················· | ···················· | ···················· | ···················· |
| G | 7156 | hCoV-19/Spain/MD-HLP-R62/2020\|EPI_ISL_530108\|2020- | ···················· | ···················· | ····················· | ···················· | ···················· | ···················· |
| G | 7157 | hCoV-19/Spain/MD-HLP-R63/2020\|EPI_ISL_530109\|2020- | ···················· | ···················· | ····················· | ···················· | ···················· | ···················· |
| G | 7158 | hCoV-19/Spain/MD-HLP-R64/2020\|EPI_ISL_530110\|2020- | ···················· | ·A·················· | ····················· | ···················· | ···················· | ···················· |
| G | 7159 | hCoV-19/Spain/MD-HLP-R65/2020\|EPI_ISL_530111\|2020- | ···················· | ···················· | ····················· | ···················· | ···················· | ···················· |
| G | 7160 | hCoV-19/Spain/MD-HLP-R67/2020\|EPI_ISL_530112\|2020- | ···················· | ···················· | ····················· | ···················· | ···················· | ···················· |
| G | 7161 | hCoV-19/Spain/MD-HLP-R68/2020\|EPI_ISL_530113\|2020- | ···················· | ···················· | ····················· | ···················· | ···················· | ···················· |
| G | 7162 | hCoV-19/Spain/MD-HLP-R81/2020\|EPI_ISL_530114\|2020- | ···················· | ···················· | ····················· | ···················· | ···················· | ···················· |
| G | 7163 | hCoV-19/Spain/MD-HLP-R82-2/2020\|EPI_ISL_530115\|202 | ···················· | ···················· | ····················· | ···················· | ···················· | ···················· |
| G | 7164 | hCoV-19/Spain/MD-HLP-R84/2020\|EPI_ISL_530116\|2020- | ···················· | ···················· | ····················· | ···················· | ···················· | ···················· |
| G | 7165 | hCoV-19/Spain/MD-HLP-R85/2020\|EPI_ISL_530117\|2020- | ···················· | ···················· | ····················· | ···················· | ···················· | ···················· |
| G | 7166 | hCoV-19/Spain/MD-HLP-R86/2020\|EPI_ISL_530118\|2020- | ···················· | ···················· | ····················· | ···················· | ···················· | ···················· |
| G | 7168 | hCoV-19/USA/WA-S2767/2020\|EPI_ISL_530124\|2020-05-1 | ···················· | ···················· | ····················· | ···················· | ···················· | ···················· |
| G | 7173 | hCoV-19/England/LIVE-A81E9/2020\|EPI_ISL_517129\|202 | ···················· | ···················· | ····················· | ···················· | ···················· | ···················· |
| G | 7174 | hCoV-19/England/LIVE-A829B/2020\|EPI_ISL_517139\|202 | ···················· | ···················· | ····················· | ···················· | ···················· | ···················· |
| G | 7175 | hCoV-19/England/LIVE-A82F5/2020\|EPI_ISL_517144\|202 | ···················· | ···················· | ····················· | ···················· | ···················· | ···················· |
| G | 7177 | hCoV-19/Australia/VIC5594/2020\|EPI_ISL_518410\|2020 | ···················· | ···················· | ····················· | ···················· | ···················· | ···················· |
| G | 7178 | hCoV-19/Australia/VIC5595/2020\|EPI_ISL_518411\|2020 | ···················· | ···················· | ····················· | ···················· | ···················· | ···················· |
| G | 7179 | hCoV-19/Australia/VIC5596/2020\|EPI_ISL_518412\|2020 | ···················· | ···················· | ····················· | ···················· | ···················· | ···················· |
| G | 7180 | hCoV-19/Australia/VIC5599/2020\|EPI_ISL_518415\|2020 | ···················· | ···················· | ····················· | ···················· | ···················· | ···················· |
| G | 7181 | hCoV-19/Australia/VIC5600/2020\|EPI_ISL_518416\|2020 | ···················· | ···················· | ····················· | ···················· | ···················· | ···················· |
| G | 7182 | hCoV-19/Australia/VIC5607/2020\|EPI_ISL_518422\|2020 | ···················· | ···················· | ····················· | ···················· | ···················· | ···················· |
| G | 7184 | hCoV-19/Australia/VIC5611/2020\|EPI_ISL_518426\|2020 | ···················· | ···················· | ····················· | ···················· | ···················· | ···················· |
| G | 7185 | hCoV-19/Australia/VIC5613/2020\|EPI_ISL_518428\|2020 | ···················· | ···················· | ····················· | ···················· | ···················· | ···················· |
| G | 7186 | hCoV-19/Australia/VIC5614/2020\|EPI_ISL_518429\|2020 | ···················· | ···················· | ····················· | ···················· | ···················· | ···················· |
| G | 7187 | hCoV-19/Australia/VIC5616/2020\|EPI_ISL_518431\|2020 | ···················· | ···················· | ····················· | ···················· | ···················· | ···················· |
| G | 7188 | hCoV-19/Australia/VIC5619/2020\|EPI_ISL_518433\|2020 | ···················· | ···················· | ····················· | ···················· | ···················· | ···················· |
| G | 7189 | hCoV-19/Australia/VIC5620/2020\|EPI_ISL_518434\|2020 | ···················· | ···················· | ····················· | ···················· | ···················· | ···················· |
| G | 7190 | hCoV-19/Australia/VIC5621/2020\|EPI_ISL_518435\|2020 | ···················· | ···················· | ····················· | ···················· | ···················· | ···················· |

| | | | | | | | | | |
|---|---|---|---|---|---|---|---|---|---|
| G | 7191 | hCoV-19/Australia/VIC5622/2020\|EPI_ISL_518436\|2020 | ···················· | ···················· | ····················· | ···················· | ···················· | ···················· | |
| G | 7192 | hCoV-19/Australia/VIC5624/2020\|EPI_ISL_518438\|2020 | ···················· | ···················· | ····················· | ···················· | ···················· | ···················· | |
| G | 7193 | hCoV-19/Australia/VIC5628/2020\|EPI_ISL_518439\|2020 | ···················· | ···················· | ····················· | ···················· | ···················· | ···················· | |
| G | 7194 | hCoV-19/Australia/VIC5629/2020\|EPI_ISL_518440\|2020 | ···················· | ···················· | ····················· | ···················· | ···················· | ···················· | |
| G | 7195 | hCoV-19/Australia/VIC5631/2020\|EPI_ISL_518442\|2020 | ···················· | ···················· | ····················· | ···················· | ···················· | ···················· | |
| G | 7196 | hCoV-19/Australia/VIC5632/2020\|EPI_ISL_518443\|2020 | ···················· | ···················· | ····················· | ···················· | ···················· | ···················· | |
| G | 7197 | hCoV-19/Australia/VIC5633/2020\|EPI_ISL_518444\|2020 | ···················· | ···················· | ····················· | ···················· | ···················· | ···················· | |
| G | 7198 | hCoV-19/Australia/VIC5634/2020\|EPI_ISL_518445\|2020 | ···················· | ···················· | ····················· | ···················· | ···················· | ···················· | |
| G | 7199 | hCoV-19/Australia/VIC5635/2020\|EPI_ISL_518446\|2020 | ···················· | ···················· | ····················· | ···················· | ···················· | ···················· | |
| G | 7200 | hCoV-19/Australia/VIC5637/2020\|EPI_ISL_518448\|2020 | ···················· | ···················· | ····················· | ···················· | ···················· | ···················· | |
| G | 7201 | hCoV-19/Australia/VIC5638/2020\|EPI_ISL_518449\|2020 | ···················· | ···················· | ····················· | ···················· | ···················· | ···················· | |
| G | 7202 | hCoV-19/Australia/VIC5641/2020\|EPI_ISL_518452\|2020 | ···················· | ···················· | ····················· | ···················· | ···················· | ···················· | |
| G | 7203 | hCoV-19/Australia/VIC5642/2020\|EPI_ISL_518453\|2020 | ···················· | ···················· | ····················· | ···················· | ···················· | ···················· | |
| G | 7204 | hCoV-19/Australia/VIC5643/2020\|EPI_ISL_518454\|2020 | ···················· | ···················· | ····················· | ···················· | ···················· | ···················· | |
| G | 7205 | hCoV-19/Australia/VIC5645/2020\|EPI_ISL_518456\|2020 | ···················· | ···················· | ····················· | ···················· | ···················· | ···················· | |
| G | 7206 | hCoV-19/Australia/VIC5646/2020\|EPI_ISL_518457\|2020 | ···················· | ···················· | ····················· | ···················· | ···················· | ···················· | |
| G | 7207 | hCoV-19/Australia/VIC5648/2020\|EPI_ISL_518459\|2020 | ···················· | ···················· | ····················· | ···················· | ···················· | ···················· | |
| G | 7208 | hCoV-19/Australia/VIC5649/2020\|EPI_ISL_518460\|2020 | ···················· | ···················· | ····················· | ···················· | ···················· | ···················· | |
| G | 7209 | hCoV-19/Australia/VIC5650/2020\|EPI_ISL_518461\|2020 | ···················· | ···················· | ····················· | ···················· | ···················· | ···················· | |
| G | 7210 | hCoV-19/Australia/VIC5652/2020\|EPI_ISL_518462\|2020 | ···················· | ···················· | ····················· | ···················· | ···················· | ···················· | |
| G | 7211 | hCoV-19/Australia/VIC5653/2020\|EPI_ISL_518463\|2020 | ···················· | ···················· | ····················· | ···················· | ···················· | ···················· | |
| G | 7212 | hCoV-19/Australia/VIC5655/2020\|EPI_ISL_518465\|2020 | ···················· | ···················· | ····················· | ···················· | ···················· | ···················· | |
| G | 7213 | hCoV-19/Australia/VIC5656/2020\|EPI_ISL_518466\|2020 | ···················· | ···················· | ····················· | ···················· | ···················· | ···················· | |
| G | 7214 | hCoV-19/Australia/VIC5657/2020\|EPI_ISL_518467\|2020 | ···················· | ···················· | ····················· | ···················· | ···················· | ···················· | |
| G | 7215 | hCoV-19/Australia/VIC5659/2020\|EPI_ISL_518469\|2020 | ···················· | ···················· | ····················· | ···················· | ···················· | ···················· | |
| G | 7216 | hCoV-19/Australia/VIC5660/2020\|EPI_ISL_518470\|2020 | ···················· | ···················· | ····················· | ···················· | ···················· | ···················· | |
| G | 7217 | hCoV-19/Australia/VIC5661/2020\|EPI_ISL_518471\|2020 | ···················· | ···················· | ····················· | ···················· | ···················· | ···················· | |
| G | 7218 | hCoV-19/Australia/VIC5662/2020\|EPI_ISL_518472\|2020 | ···················· | ···················· | ····················· | ···················· | ···················· | ···················· | |
| G | 7219 | hCoV-19/Australia/VIC5665/2020\|EPI_ISL_518473\|2020 | ···················· | ···················· | ····················· | ···················· | ···················· | ···················· | |
| G | 7220 | hCoV-19/Australia/VIC5667/2020\|EPI_ISL_518474\|2020 | ···················· | ···················· | ····················· | ···················· | ···················· | ···················· | |
| G | 7221 | hCoV-19/Australia/VIC5668/2020\|EPI_ISL_518475\|2020 | ···················· | ···················· | ····················· | ···················· | ···················· | ···················· | |
| G | 7222 | hCoV-19/Australia/VIC5669/2020\|EPI_ISL_518476\|2020 | ···················· | ···················· | ····················· | ···················· | ···················· | ···················· | |
| G | 7223 | hCoV-19/Australia/VIC5670/2020\|EPI_ISL_518477\|2020 | ···················· | ···················· | ····················· | ···················· | ···················· | ···················· | |
| G | 7224 | hCoV-19/Australia/VIC5672/2020\|EPI_ISL_518479\|2020 | ···················· | ···················· | ····················· | ···················· | ···················· | ···················· | |
| G | 7225 | hCoV-19/Australia/VIC4444/2020\|EPI_ISL_519740\|2020 | ···················· | ···················· | ····················· | ···················· | ···················· | ···················· | |
| G | 7226 | hCoV-19/Australia/VIC4445/2020\|EPI_ISL_519741\|2020 | ···················· | ···················· | ····················· | ···················· | ···················· | ···················· | |
| G | 7227 | hCoV-19/Australia/VIC4446/2020\|EPI_ISL_519742\|2020 | ···················· | ···················· | ····················· | ···················· | ···················· | ···················· | |
| G | 7228 | hCoV-19/Australia/VIC4447/2020\|EPI_ISL_519743\|2020 | ···················· | ···················· | ····················· | ···················· | ···················· | ···················· | |
| G | 7229 | hCoV-19/Australia/VIC4448/2020\|EPI_ISL_519744\|2020 | ···················· | ···················· | ····················· | ···················· | ···················· | ···················· | |
| G | 7230 | hCoV-19/Australia/VIC4449/2020\|EPI_ISL_519745\|2020 | ···················· | ···················· | ····················· | ···················· | ···················· | ···················· | |
| G | 7231 | hCoV-19/Australia/VIC4450/2020\|EPI_ISL_519746\|2020 | ···················· | ···················· | ····················· | ···················· | ···················· | ···················· | |
| G | 7232 | hCoV-19/Australia/VIC4451/2020\|EPI_ISL_519747\|2020 | ···················· | ···················· | ····················· | ···················· | ···················· | ···················· | |
| G | 7233 | hCoV-19/Australia/VIC4452/2020\|EPI_ISL_519748\|2020 | ···················· | ···················· | ····················· | ···················· | ···················· | ···················· | |
| G | 7234 | hCoV-19/Australia/VIC4453/2020\|EPI_ISL_519749\|2020 | ···················· | ···················· | ····················· | ···················· | ···················· | ···················· | |
| G | 7235 | hCoV-19/Australia/VIC4454/2020\|EPI_ISL_519750\|2020 | ···················· | ···················· | ····················· | ···················· | ···················· | ···················· | |
| G | 7236 | hCoV-19/Australia/VIC4455/2020\|EPI_ISL_519751\|2020 | ···················· | ···················· | ····················· | ···················· | ···················· | ···················· | |
| G | 7237 | hCoV-19/Australia/VIC4456/2020\|EPI_ISL_519752\|2020 | ···················· | ···················· | ····················· | ···················· | ···················· | ···················· | |
| G | 7238 | hCoV-19/Australia/VIC4457/2020\|EPI_ISL_519753\|2020 | ···················· | ···················· | ····················· | ···················· | ···················· | ···················· | |
| G | 7239 | hCoV-19/Australia/VIC4458/2020\|EPI_ISL_519754\|2020 | ···················· | ···················· | ····················· | ···················· | ···················· | ···················· | |
| G | 7240 | hCoV-19/Australia/VIC4459/2020\|EPI_ISL_519755\|2020 | ···················· | ···················· | ····················· | ···················· | ···················· | ···················· | |
| G | 7241 | hCoV-19/Australia/VIC4461/2020\|EPI_ISL_519757\|2020 | ···················· | ···················· | ····················· | ···················· | ···················· | ···················· | |
| G | 7242 | hCoV-19/Australia/VIC4462/2020\|EPI_ISL_519758\|2020 | ···················· | ···················· | ····················· | ···················· | ···················· | ···················· | |
| G | 7243 | hCoV-19/Australia/VIC4463/2020\|EPI_ISL_519759\|2020 | ···················· | ···················· | ····················· | ···················· | ···················· | ···················· | |
| G | 7244 | hCoV-19/Australia/VIC4464/2020\|EPI_ISL_519760\|2020 | ···················· | ···················· | ····················· | ···················· | ···················· | ···················· | |
| G | 7245 | hCoV-19/Australia/VIC4465/2020\|EPI_ISL_519761\|2020 | ···················· | ···················· | ····················· | ···················· | ···················· | ···················· | |
| G | 7246 | hCoV-19/Australia/VIC4466/2020\|EPI_ISL_519762\|2020 | ···················· | ···················· | ····················· | ···················· | ···················· | ···················· | |
| G | 7247 | hCoV-19/Australia/VIC4467/2020\|EPI_ISL_519763\|2020 | ··················T· | ···················· | ····················· | ···················· | ···················· | ···················· | 1 |
| G | 7248 | hCoV-19/Australia/VIC4468/2020\|EPI_ISL_519764\|2020 | ···················· | ···················· | ····················· | ···················· | ···················· | ···················· | |
| G | 7249 | hCoV-19/Australia/VIC4469/2020\|EPI_ISL_519765\|2020 | ···················· | ···················· | ····················· | ···················· | ···················· | ···················· | |
| G | 7250 | hCoV-19/Australia/VIC4470/2020\|EPI_ISL_519766\|2020 | ···················· | ···················· | ····················· | ···················· | ···················· | ···················· | |
| G | 7251 | hCoV-19/Australia/VIC4471/2020\|EPI_ISL_519767\|2020 | ···················· | ···················· | ····················· | ···················· | ···················· | ···················· | |
| G | 7252 | hCoV-19/Australia/VIC4472/2020\|EPI_ISL_519768\|2020 | ···················· | ···················· | ····················· | ···················· | ···················· | ···················· | |
| G | 7253 | hCoV-19/Australia/VIC4473/2020\|EPI_ISL_519769\|2020 | ···················· | ···················· | ····················· | ···················· | ···················· | ···················· | |
| G | 7254 | hCoV-19/Australia/VIC4474/2020\|EPI_ISL_519770\|2020 | ···················· | ···················· | ····················· | ···················· | ···················· | ···················· | |
| G | 7255 | hCoV-19/Australia/VIC4475/2020\|EPI_ISL_519771\|2020 | ···················· | ···················· | ····················· | ···················· | ···················· | ···················· | |
| G | 7256 | hCoV-19/Australia/VIC4476/2020\|EPI_ISL_519772\|2020 | ···················· | ···················· | ····················· | ···················· | ···················· | ···················· | |
| G | 7257 | hCoV-19/Australia/VIC4477/2020\|EPI_ISL_519773\|2020 | ···················· | ···················· | ····················· | ···················· | ···················· | ···················· | |
| G | 7258 | hCoV-19/Australia/VIC4478/2020\|EPI_ISL_519774\|2020 | ···················· | ···················· | ····················· | ···················· | ···················· | ···················· | |
| G | 7259 | hCoV-19/Australia/VIC4479/2020\|EPI_ISL_519775\|2020 | ···················· | ···················· | ····················· | ···················· | ···················· | ···················· | |
| G | 7260 | hCoV-19/Australia/VIC4480/2020\|EPI_ISL_519776\|2020 | ···················· | ···················· | ····················· | ···················· | ···················· | ···················· | |
| G | 7261 | hCoV-19/Australia/VIC4481/2020\|EPI_ISL_519777\|2020 | ···················· | ···················· | ····················· | ···················· | ···················· | ···················· | |
| G | 7262 | hCoV-19/Australia/VIC4482/2020\|EPI_ISL_519778\|2020 | ···················· | ···················· | ····················· | ···················· | ···················· | ···················· | |
| G | 7263 | hCoV-19/Australia/VIC4483/2020\|EPI_ISL_519779\|2020 | ···················· | ···················· | ····················· | ···················· | ···················· | ···················· | |
| G | 7264 | hCoV-19/Australia/VIC4484/2020\|EPI_ISL_519780\|2020 | ···················· | ···················· | ····················· | ···················· | ···················· | ···················· | |
| G | 7265 | hCoV-19/Australia/VIC4485/2020\|EPI_ISL_519781\|2020 | ···················· | ···················· | ····················· | ···················· | ···················· | ···················· | |
| G | 7266 | hCoV-19/Australia/VIC4487/2020\|EPI_ISL_519783\|2020 | ···················· | ···················· | ····················· | ···················· | ···················· | ···················· | |
| G | 7267 | hCoV-19/Australia/VIC4488/2020\|EPI_ISL_519784\|2020 | ···················· | ···················· | ····················· | ···················· | ···················· | ···················· | |
| G | 7268 | hCoV-19/Australia/VIC4982/2020\|EPI_ISL_519786\|2020 | ···················· | ···················· | ····················· | ···················· | ···················· | ···················· | |
| G | 7269 | hCoV-19/Australia/VIC4983/2020\|EPI_ISL_519787\|2020 | ···················· | ···················· | ····················· | ···················· | ···················· | ···················· | |
| G | 7270 | hCoV-19/Australia/VIC4984/2020\|EPI_ISL_519788\|2020 | ···················· | ···················· | ····················· | ···················· | ···················· | ···················· | |

| | | | | | | | | |
|---|---|---|---|---|---|---|---|---|
| G | 7271 hCoV-19/Australia/VIC4985/2020\|EPI_ISL_519789\|2020 | ···················· | ···················· | ···················· | ···················· | ···················· | ···················· | |
| G | 7272 hCoV-19/Australia/VIC4986/2020\|EPI_ISL_519790\|2020 | ···················· | ···················· | ···················· | ···················· | ···················· | ···················· | |
| G | 7273 hCoV-19/Australia/VIC4987/2020\|EPI_ISL_519791\|2020 | ···················· | ···················· | ···················· | ···················· | ···················· | ···················· | |
| G | 7274 hCoV-19/Australia/VIC4988/2020\|EPI_ISL_519792\|2020 | ···················· | ···················· | ···················· | ···················· | ···················· | ···················· | |
| G | 7275 hCoV-19/Australia/VIC4989/2020\|EPI_ISL_519793\|2020 | ···················· | ···················· | ···················· | ···················· | ···················· | ···················· | |
| G | 7276 hCoV-19/Australia/VIC4990/2020\|EPI_ISL_519794\|2020 | ···················· | ···················· | ···················· | ···················· | ···················· | ···················· | |
| G | 7277 hCoV-19/Australia/VIC4991/2020\|EPI_ISL_519795\|2020 | ···················· | ···················· | ···················· | ···················· | ···················· | ···················· | |
| G | 7278 hCoV-19/Australia/VIC4992/2020\|EPI_ISL_519796\|2020 | ···················· | ···················· | ···················· | ···················· | ···················· | ···················· | |
| G | 7279 hCoV-19/Australia/VIC4993/2020\|EPI_ISL_519797\|2020 | ···················· | ···················· | ···················· | ···················· | ···················· | ···················· | |
| G | 7280 hCoV-19/Australia/VIC4994/2020\|EPI_ISL_519798\|2020 | ···················· | ···················· | ···················· | ···················· | ···················· | ···················· | |
| G | 7281 hCoV-19/Australia/VIC4995/2020\|EPI_ISL_519799\|2020 | ···················· | ···················· | ···················· | ···················· | ···················· | ···················· | |
| G | 7282 hCoV-19/United Arab Emirates/L920632111/2020\|EPI_I | ···················· | ···················· | ···················· | ···················· | ···················· | ···················· | |
| G | 7283 hCoV-19/United Arab Emirates/L920632662/2020\|EPI_I | ···················· | ···················· | ···················· | ···················· | ···················· | ···················· | |
| G | 7284 hCoV-19/United Arab Emirates/L920632696/2020\|EPI_I | ···················· | ···················· | ···················· | ···················· | ···················· | ···················· | |
| G | 7285 hCoV-19/United Arab Emirates/L920633045/2020\|EPI_I | ···················· | ···················· | ···················· | ···················· | ···················· | ···················· | |
| G | 7286 hCoV-19/United Arab Emirates/L920633058/2020\|EPI_I | ···················· | ···················· | ···················· | ···················· | ···················· | ···················· | |
| G | 7287 hCoV-19/United Arab Emirates/L920633304/2020\|EPI_I | ···················· | ···················· | ···················· | ···················· | ···················· | ···················· | |
| G | 7288 hCoV-19/United Arab Emirates/L920633851/2020\|EPI_I | ···················· | ···················· | ···················· | ···················· | ···················· | ···················· | |
| G | 7289 hCoV-19/United Arab Emirates/L920634181/2020\|EPI_I | ···················· | ···················· | ···················· | ···················· | ···················· | ···················· | |
| G | 7290 hCoV-19/United Arab Emirates/L920634731/2020\|EPI_I | ···················· | ···················· | ···················· | ···················· | ···················· | ···················· | |
| G | 7291 hCoV-19/United Arab Emirates/L920638060/2020\|EPI_I | ···················· | ···················· | ···················· | ···················· | ···················· | ···················· | |
| G | 7292 hCoV-19/United Arab Emirates/L920639763/2020\|EPI_I | ···················· | ···················· | ···················· | ···················· | ···················· | ···················· | |
| G | 7293 hCoV-19/United Arab Emirates/L920643384/2020\|EPI_I | ···················· | ···················· | ···················· | ···················· | ···················· | ···················· | |
| G | 7294 hCoV-19/United Arab Emirates/L920647384/2020\|EPI_I | ···················· | ···················· | ···················· | ···················· | ···················· | ···················· | |
| G | 7295 hCoV-19/United Arab Emirates/L920647430/2020\|EPI_I | ···················· | ···················· | ···················· | ···················· | ···················· | ···················· | |
| G | 7296 hCoV-19/United Arab Emirates/L920663561/2020\|EPI_I | ···················· | ···················· | ···················· | ···················· | ···················· | ···················· | |
| G | 7297 hCoV-19/United Arab Emirates/L920667218/2020\|EPI_I | ···················· | ···················· | ···················· | ···················· | ···················· | ···················· | |
| G | 7298 hCoV-19/United Arab Emirates/L920667894/2020\|EPI_I | ···················· | ···················· | ···················· | ···················· | ···················· | ···················· | |
| G | 7299 hCoV-19/United Arab Emirates/L920673013/2020\|EPI_I | ···················· | ···················· | ···················· | ···················· | ···················· | ···················· | |
| G | 7300 hCoV-19/United Arab Emirates/L920689030/2020\|EPI_I | ···················· | ···················· | ···················· | ···················· | ···················· | ···················· | |
| G | 7301 hCoV-19/United Arab Emirates/L920691421/2020\|EPI_I | ···················· | ···················· | ···················· | ···················· | ···················· | ···················· | |
| G | 7302 hCoV-19/United Arab Emirates/L920702211/2020\|EPI_I | ···················· | ···················· | ···················· | ···················· | ···················· | ···················· | |
| G | 7303 hCoV-19/United Arab Emirates/L920707982/2020\|EPI_I | ···················· | ···················· | ···················· | ···················· | ···················· | ···················· | |
| G | 7304 hCoV-19/United Arab Emirates/L920716500/2020\|EPI_I | ···················· | ···················· | ···················· | ···················· | ···················· | ···················· | |
| G | 7305 hCoV-19/United Arab Emirates/L9584510/2020\|EPI_ISL | ···················· | ···················· | ···················· | ···················· | ···················· | ···················· | |
| G | 7306 hCoV-19/United Arab Emirates/L9986375/2020\|EPI_ISL | ···················· | ···················· | ···················· | ···················· | ···················· | ···················· | |
| G | 7307 hCoV-19/Australia/VIC3031/2020\|EPI_ISL_520745\|2020 | ···················· | ···················· | ···················· | ···················· | ···················· | ···················· | |
| G | 7308 hCoV-19/Australia/VIC3032/2020\|EPI_ISL_520746\|2020 | ···················· | ···················· | ···················· | ···················· | ···················· | ···················· | |
| G | 7309 hCoV-19/Australia/VIC3033/2020\|EPI_ISL_520747\|2020 | ···················· | ···················· | ···················· | ···················· | ···················· | ···················· | |
| G | 7310 hCoV-19/Australia/VIC3035/2020\|EPI_ISL_520748\|2020 | ···················· | ···················· | ···················· | ···················· | ···················· | ···················· | |
| G | 7311 hCoV-19/Australia/VIC3036/2020\|EPI_ISL_520749\|2020 | ···················· | ···················· | ···················· | ···················· | ···················· | ···················· | |
| G | 7312 hCoV-19/Australia/VIC3037/2020\|EPI_ISL_520750\|2020 | ···················· | ···················· | ···················· | ···················· | ···················· | ···················· | |
| G | 7313 hCoV-19/Australia/VIC3039/2020\|EPI_ISL_520752\|2020 | ···················· | ···················· | ···················· | ···················· | ···················· | ···················· | |
| G | 7314 hCoV-19/Australia/VIC3040/2020\|EPI_ISL_520753\|2020 | ···················· | ···················· | ···················· | ···················· | ···················· | ···················· | |
| G | 7315 hCoV-19/Australia/VIC3041/2020\|EPI_ISL_520754\|2020 | ···················· | ···················· | ···················· | ···················· | ···················· | ···················· | |
| G | 7316 hCoV-19/Australia/VIC3042/2020\|EPI_ISL_520755\|2020 | ···················· | ···················· | ···················· | ···················· | ···················· | ···················· | |
| G | 7317 hCoV-19/Australia/VIC3043/2020\|EPI_ISL_520756\|2020 | ···················· | ···················· | ···················· | ···················· | ···················· | ···················· | |
| G | 7318 hCoV-19/Australia/VIC3044/2020\|EPI_ISL_520757\|2020 | ···················· | ···················· | ···················· | ···················· | ···················· | ···················· | |
| G | 7319 hCoV-19/Australia/VIC3045/2020\|EPI_ISL_520758\|2020 | ···················· | ···················· | ···················· | ···················· | ···················· | ···················· | |
| G | 7320 hCoV-19/Australia/VIC3046/2020\|EPI_ISL_520759\|2020 | ···················· | ···················· | ···················· | ···················· | ···················· | ···················· | |
| G | 7321 hCoV-19/Australia/VIC3047/2020\|EPI_ISL_520760\|2020 | ···················· | ···················· | ···················· | ···················· | ···················· | ···················· | |
| G | 7323 hCoV-19/Australia/VIC3049/2020\|EPI_ISL_520762\|2020 | ···················· | ···················· | ···················· | ···················· | ···················· | ···················· | |
| G | 7324 hCoV-19/Australia/VIC3058/2020\|EPI_ISL_520765\|2020 | ···················· | ···················· | ···················· | ···················· | ···················· | ···················· | |
| G | 7325 hCoV-19/Australia/VIC3059/2020\|EPI_ISL_520766\|2020 | ···················· | ···················· | ···················· | ···················· | ···················· | ···················· | |
| G | 7327 hCoV-19/Australia/VIC3061/2020\|EPI_ISL_520768\|2020 | ···················· | ···················· | ···················· | ···················· | NNN················· | ···················· | 1 |
| G | 7328 hCoV-19/Australia/VIC3062/2020\|EPI_ISL_520769\|2020 | ···················· | ···················· | ···················· | ···················· | ···················· | ···················· | |
| G | 7329 hCoV-19/Australia/VIC3063/2020\|EPI_ISL_520770\|2020 | ···················· | ···················· | ···················· | ···················· | ···················· | ···················· | |
| G | 7331 hCoV-19/Australia/VIC3065/2020\|EPI_ISL_520772\|2020 | ···················· | ···················· | ···················· | ···················· | ···················· | ···················· | |
| G | 7333 hCoV-19/Australia/VIC3067/2020\|EPI_ISL_520774\|2020 | ···················· | ···················· | ···················· | ···················· | ···················· | ···················· | |
| G | 7334 hCoV-19/Australia/VIC3069/2020\|EPI_ISL_520776\|2020 | ···················· | ···················· | ···················· | ···················· | ···················· | ···················· | |
| G | 7336 hCoV-19/Australia/VIC3071/2020\|EPI_ISL_520778\|2020 | ···················· | ···················· | ···················· | ···················· | ···················· | ···················· | |
| G | 7337 hCoV-19/Australia/VIC3072/2020\|EPI_ISL_520779\|2020 | ···················· | ···················· | ···················· | ···················· | ···················· | ···················· | |
| G | 7338 hCoV-19/Australia/VIC3073/2020\|EPI_ISL_520780\|2020 | ···················· | ···················· | ···················· | ···················· | ···················· | ···················· | |
| G | 7339 hCoV-19/Australia/VIC3074/2020\|EPI_ISL_520781\|2020 | ···················· | ···················· | ···················· | ···················· | ···················· | ···················· | |
| G | 7340 hCoV-19/Australia/VIC3075/2020\|EPI_ISL_520782\|2020 | ···················· | ···················· | ···················· | ···················· | ···················· | ···················· | |
| G | 7342 hCoV-19/Australia/VIC3077/2020\|EPI_ISL_520784\|2020 | ···················· | ···················· | ···················· | ···················· | ···················· | ···················· | |
| G | 7343 hCoV-19/Australia/VIC3078/2020\|EPI_ISL_520785\|2020 | ···················· | ···················· | ···················· | ···················· | ···················· | ···················· | |
| G | 7344 hCoV-19/Australia/VIC3079/2020\|EPI_ISL_520786\|2020 | ···················· | ···················· | ···················· | ···················· | ···················· | ···················· | |
| G | 7345 hCoV-19/Australia/VIC3080/2020\|EPI_ISL_520787\|2020 | ···················· | ···················· | ···················· | ···················· | ···················· | ···················· | |
| G | 7346 hCoV-19/Australia/VIC3081/2020\|EPI_ISL_520788\|2020 | ···················· | ···················· | ···················· | ···················· | ···················· | ···················· | |
| G | 7347 hCoV-19/Australia/VIC3082/2020\|EPI_ISL_520789\|2020 | ···················· | ···················· | ···················· | ···················· | ···················· | ···················· | |
| G | 7348 hCoV-19/Australia/VIC3083/2020\|EPI_ISL_520790\|2020 | ···················· | ···················· | ···················· | ···················· | ···················· | ···················· | |
| G | 7349 hCoV-19/Australia/VIC3084/2020\|EPI_ISL_520791\|2020 | ···················· | ···················· | ···················· | ···················· | ···················· | ···················· | |
| G | 7351 hCoV-19/Australia/VIC3086/2020\|EPI_ISL_520793\|2020 | ···················· | ···················· | ···················· | ···················· | ···················· | ···················· | |
| G | 7352 hCoV-19/Australia/VIC3087/2020\|EPI_ISL_520794\|2020 | ···················· | ···················· | ···················· | ···················· | ···················· | ···················· | |
| G | 7353 hCoV-19/Australia/VIC3088/2020\|EPI_ISL_520795\|2020 | ···················· | ···················· | ···················· | ···················· | ···················· | ···················· | |
| G | 7354 hCoV-19/Australia/VIC3089/2020\|EPI_ISL_520796\|2020 | ···················· | ···················· | ···················· | ···················· | ···················· | ···················· | |
| G | 7355 hCoV-19/Australia/VIC3321/2020\|EPI_ISL_521000\|2020 | ···················· | ···················· | ···················· | ···················· | ···················· | ···················· | |
| G | 7356 hCoV-19/Australia/VIC3322/2020\|EPI_ISL_521001\|2020 | ···················· | ···················· | ···················· | ···················· | ···················· | ···················· | |
| G | 7357 hCoV-19/Australia/VIC3323/2020\|EPI_ISL_521002\|2020 | ···················· | ···················· | ···················· | ···················· | ···················· | ···················· | |

| | | | | | | | | |
|---|---|---|---|---|---|---|---|---|
| G | 7358 hCoV-19/Australia/VIC3324/2020\|EPI_ISL_521003\|2020 | ···················· | ···················· | ···················· | ···················· | ···················· | ···················· | |
| G | 7359 hCoV-19/Australia/VIC3325/2020\|EPI_ISL_521004\|2020 | ···················· | ···················· | ···················· | ···················· | ···················· | ···················· | |
| G | 7360 hCoV-19/Australia/VIC3326/2020\|EPI_ISL_521005\|2020 | ···················· | ···················· | ···················· | ···················· | ···················· | ···················· | |
| G | 7361 hCoV-19/Australia/VIC3327/2020\|EPI_ISL_521006\|2020 | ···················· | ···················· | ···················· | ···················· | ···················· | ···················· | |
| G | 7362 hCoV-19/Australia/VIC3328/2020\|EPI_ISL_521007\|2020 | ···················· | ···················· | ···················· | ···················· | ···················· | ···················· | |
| G | 7363 hCoV-19/Australia/VIC3329/2020\|EPI_ISL_521008\|2020 | ···················· | ···················· | ···················· | ···················· | ···················· | ···················· | |
| G | 7364 hCoV-19/Australia/VIC3330/2020\|EPI_ISL_521009\|2020 | ···················· | ···················· | ···················· | ···················· | ···················· | ···················· | |
| G | 7365 hCoV-19/Australia/VIC3331/2020\|EPI_ISL_521010\|2020 | ···················· | ···················· | ···················· | ···················· | ···················· | ···················· | |
| G | 7366 hCoV-19/Australia/VIC3332/2020\|EPI_ISL_521011\|2020 | ···················· | ···················· | ···················· | ···················· | ···················· | ···················· | |
| G | 7367 hCoV-19/Australia/VIC3333/2020\|EPI_ISL_521012\|2020 | ···················· | ···················· | ···················· | ···················· | ···················· | ···················· | |
| G | 7368 hCoV-19/Australia/VIC3334/2020\|EPI_ISL_521013\|2020 | ···················· | ···················· | ···················· | ···················· | ···················· | ···················· | |
| G | 7369 hCoV-19/Australia/VIC3335/2020\|EPI_ISL_521014\|2020 | ···················· | ···················· | ···················· | ···················· | ···················· | ···················· | |
| G | 7370 hCoV-19/Australia/VIC3336/2020\|EPI_ISL_521015\|2020 | ···················· | ···················· | ···················· | ···················· | ···················· | ···················· | |
| G | 7371 hCoV-19/Australia/VIC3337/2020\|EPI_ISL_521016\|2020 | ···················· | ···················· | ···················· | ···················· | ···················· | ···················· | |
| G | 7372 hCoV-19/USA/UT-04276/2020\|EPI_ISL_522331\|2020-05-1 | ···················· | ···················· | ···················· | ···················· | ···················· | ···················· | |
| G | 7373 hCoV-19/USA/UT-04288/2020\|EPI_ISL_522336\|2020-05-1 | ···················· | ···················· | ···················· | ···················· | ···················· | ···················· | |
| G | 7374 hCoV-19/USA/UT-04291/2020\|EPI_ISL_522337\|2020-05-1 | ···················· | ···················· | ···················· | ···················· | ···················· | ···················· | |
| G | 7375 hCoV-19/USA/UT-04295/2020\|EPI_ISL_522340\|2020-05-1 | ···················· | ·A·················· | ···················· | ···················· | ···················· | ···················· | |
| G | 7376 hCoV-19/USA/UT-04480/2020\|EPI_ISL_522341\|2020-05-2 | ···················· | ···················· | ···················· | ···················· | ···················· | ···················· | |
| G | 7377 hCoV-19/USA/UT-04499/2020\|EPI_ISL_522343\|2020-05-2 | ···················· | ···················· | ···················· | ···················· | ···················· | ···················· | |
| G | 7378 hCoV-19/Netherlands/ZH-EMC-308/2020\|EPI_ISL_523650 | ···················· | ···················· | ···················· | ···················· | ···················· | ···················· | |
| G | 7379 hCoV-19/Netherlands/ZH-EMC-309/2020\|EPI_ISL_523651 | ···················· | ···················· | ···················· | ···················· | ···················· | ···················· | |
| G | 7380 hCoV-19/Netherlands/ZH-EMC-310/2020\|EPI_ISL_523652 | ···················· | ···················· | ···················· | ···················· | ···················· | ···················· | |
| G | 7381 hCoV-19/Netherlands/ZH-EMC-312/2020\|EPI_ISL_523654 | ···················· | ···················· | ···················· | ···················· | ···················· | ···················· | |
| G | 7382 hCoV-19/Netherlands/ZH-EMC-315/2020\|EPI_ISL_523657 | ···················· | ···················· | ···················· | ···················· | ···················· | ···················· | |
| G | 7383 hCoV-19/Netherlands/ZH-EMC-316/2020\|EPI_ISL_523658 | ···················· | ···················· | ···················· | ···················· | ···················· | ···················· | |
| G | 7384 hCoV-19/Netherlands/ZH-EMC-317/2020\|EPI_ISL_523659 | ···················· | ···················· | ···················· | ···················· | ···················· | ···················· | |
| G | 7385 hCoV-19/Netherlands/ZH-EMC-319/2020\|EPI_ISL_523660 | ···················· | ···················· | ···················· | ···················· | ···················· | ···················· | |
| G | 7386 hCoV-19/Netherlands/ZH-EMC-320/2020\|EPI_ISL_523661 | ···················· | ···················· | ···················· | ···················· | ···················· | ···················· | |
| G | 7387 hCoV-19/Netherlands/ZH-EMC-321/2020\|EPI_ISL_523662 | ···················· | ···················· | ···················· | ···················· | ···················· | ···················· | |
| G | 7388 hCoV-19/Netherlands/ZH-EMC-322/2020\|EPI_ISL_523663 | ···················· | ···················· | ···················· | ···················· | ···················· | ···················· | |
| G | 7389 hCoV-19/Netherlands/ZH-EMC-323/2020\|EPI_ISL_523664 | ···················· | ···················· | ···················· | ···················· | ···················· | ···················· | |
| G | 7390 hCoV-19/Netherlands/ZH-EMC-324/2020\|EPI_ISL_523665 | ···················· | ···················· | ···················· | ···················· | ···················· | ···················· | |
| G | 7391 hCoV-19/Netherlands/ZH-EMC-325/2020\|EPI_ISL_523666 | ···················· | ···················· | ···················· | ···················· | ···················· | ···················· | |
| G | 7392 hCoV-19/Netherlands/ZH-EMC-326/2020\|EPI_ISL_523667 | ···················· | ···················· | ···················· | ···················· | ···················· | ···················· | |
| G | 7393 hCoV-19/Netherlands/ZH-EMC-327/2020\|EPI_ISL_523668 | ···················· | ···················· | ···················· | ···················· | ···················· | ···················· | |
| G | 7394 hCoV-19/Netherlands/ZH-EMC-328/2020\|EPI_ISL_523669 | ···················· | ···················· | ···················· | ···················· | ···················· | ···················· | |
| G | 7395 hCoV-19/Netherlands/ZH-EMC-329/2020\|EPI_ISL_523670 | ···················· | ···················· | ···················· | ···················· | ···················· | ···················· | |
| G | 7396 hCoV-19/Netherlands/ZH-EMC-330/2020\|EPI_ISL_523671 | ···················· | ···················· | ···················· | ···················· | ···················· | ···················· | |
| G | 7397 hCoV-19/Netherlands/ZH-EMC-331/2020\|EPI_ISL_523672 | ···················· | ···················· | ···················· | ···················· | ···················· | ···················· | |
| G | 7398 hCoV-19/Netherlands/ZH-EMC-332/2020\|EPI_ISL_523673 | ···················· | ···················· | ···················· | ···················· | ···················· | ···················· | |
| G | 7399 hCoV-19/Netherlands/ZH-EMC-333/2020\|EPI_ISL_523674 | ···················· | ···················· | ···················· | ···················· | ···················· | ···················· | |
| G | 7400 hCoV-19/Netherlands/ZH-EMC-334/2020\|EPI_ISL_523675 | ···················· | ···················· | ···················· | ···················· | ···················· | ···················· | |
| G | 7401 hCoV-19/Netherlands/ZH-EMC-335/2020\|EPI_ISL_523676 | ···················· | ···················· | ···················· | ···················· | ···················· | ···················· | |
| G | 7402 hCoV-19/Netherlands/ZH-EMC-336/2020\|EPI_ISL_523677 | ···················· | ···················· | ···················· | ···················· | ···················· | ···················· | |
| G | 7403 hCoV-19/Netherlands/ZH-EMC-337/2020\|EPI_ISL_523678 | ···················· | ···················· | ···················· | ···················· | ···················· | ···················· | |
| G | 7404 hCoV-19/USA/UT-UPHL-2008863/2020\|EPI_ISL_524980\|20 | ···················· | ···················· | ···················· | ···················· | ···················· | ···················· | |
| G | 7405 hCoV-19/USA/UT-UPHL-2008867/2020\|EPI_ISL_524984\|20 | ···················· | ···················· | ···················· | ···················· | ···················· | ···················· | |
| G | 7406 hCoV-19/USA/UT-UPHL-2008870/2020\|EPI_ISL_524985\|20 | ···················· | ···················· | ···················· | ···················· | ···················· | ···················· | |
| G | 7407 hCoV-19/USA/UT-UPHL-2008871/2020\|EPI_ISL_524986\|20 | ···················· | ···················· | ···················· | ···················· | ···················· | ···················· | |
| G | 7408 hCoV-19/USA/UT-UPHL-2008874/2020\|EPI_ISL_524988\|20 | ···················· | ···················· | ···················· | ···················· | ···················· | ···················· | |
| G | 7409 hCoV-19/USA/UT-UPHL-2008876/2020\|EPI_ISL_524990\|20 | ···················· | ···················· | ···················· | ···················· | ···················· | ···················· | |
| G | 7410 hCoV-19/USA/UT-UPHL-2008877/2020\|EPI_ISL_524991\|20 | ···················· | ···················· | ···················· | ···················· | ···················· | ···················· | |
| G | 7411 hCoV-19/USA/UT-UPHL-2008878/2020\|EPI_ISL_524992\|20 | ···················· | ···················· | ···················· | ···················· | ···················· | ···················· | |
| G | 7412 hCoV-19/USA/UT-UPHL-2008879/2020\|EPI_ISL_524993\|20 | ···················· | ···················· | ···················· | ···················· | ···················· | ···················· | |
| G | 7413 hCoV-19/USA/UT-UPHL-2008880/2020\|EPI_ISL_524994\|20 | ···················· | ···················· | ···················· | ···················· | ···················· | ···················· | |
| G | 7414 hCoV-19/USA/UT-UPHL-2008881/2020\|EPI_ISL_524995\|20 | ···················· | ···················· | ···················· | ···················· | ···················· | ···················· | |
| G | 7415 hCoV-19/USA/UT-UPHL-2008882/2020\|EPI_ISL_524996\|20 | ···················· | ···················· | ···················· | ···················· | ···················· | ···················· | |
| G | 7416 hCoV-19/USA/UT-UPHL-2008883/2020\|EPI_ISL_524997\|20 | ···················· | ···················· | ···················· | ···················· | ···················· | ···················· | |
| G | 7417 hCoV-19/USA/UT-UPHL-2008885/2020\|EPI_ISL_524999\|20 | ···················· | ···················· | ···················· | ···················· | ···················· | ···················· | |
| G | 7418 hCoV-19/USA/SC-DHEC-0591/2020\|EPI_ISL_529100\|2020- | ···················· | ···················· | ···················· | ···················· | ···················· | ···················· | |
| G | 7419 hCoV-19/USA/SC-DHEC-0653/2020\|EPI_ISL_529101\|2020- | ···················· | ···················· | ···················· | ···················· | ···················· | ···················· | |
| G | 7420 hCoV-19/USA/SC-DHEC-0657/2020\|EPI_ISL_529103\|2020- | ···················· | ···················· | ···················· | ···················· | ···················· | ···················· | |
| G | 7421 hCoV-19/USA/SC-DHEC-0699/2020\|EPI_ISL_529105\|2020- | ···················· | ·········A·········· | ···················· | ···················· | ···················· | ···················· | |
| G | 7422 hCoV-19/USA/SC-DHEC-0708/2020\|EPI_ISL_529106\|2020- | ···················· | ···················· | ···················· | ···················· | ···················· | ···················· | |
| G | 7423 hCoV-19/USA/SC-DHEC-0709/2020\|EPI_ISL_529107\|2020- | ···················· | ···················· | ···················· | ···················· | ···················· | ···················· | |
| G | 7424 hCoV-19/USA/SC-DHEC-0710/2020\|EPI_ISL_529108\|2020- | ···················· | ···················· | ···················· | ···················· | ···················· | ···················· | |
| G | 7425 hCoV-19/USA/FL-BPHL-0910/2020\|EPI_ISL_517800\|2020- | ···················· | ···················· | ···················· | ···················· | NNN················· | ···················· | 1 |
| G | 7426 hCoV-19/USA/FL-BPHL-0911/2020\|EPI_ISL_517801\|2020- | ···················· | ···················· | ···················· | ···················· | ···················· | ···················· | |
| G | 7427 hCoV-19/USA/FL-BPHL-0912/2020\|EPI_ISL_517802\|2020- | ···················· | ·············A······ | ···················· | ···················· | ···················· | ···················· | |
| G | 7428 hCoV-19/USA/FL-BPHL-0913/2020\|EPI_ISL_517803\|2020- | ···················· | ···················· | ···················· | ···················· | ···················· | ···················· | |
| G | 7430 hCoV-19/USA/FL-BPHL-0915/2020\|EPI_ISL_517805\|2020- | ···················· | ···················· | ···················· | ···················· | ···················· | ···················· | |
| G | 7432 hCoV-19/USA/FL-BPHL-0917/2020\|EPI_ISL_517807\|2020- | ···················· | ···················· | ···················· | ···················· | ···················· | ···················· | |
| G | 7433 hCoV-19/USA/FL-BPHL-0918/2020\|EPI_ISL_517808\|2020- | ···················· | ···················· | ···················· | ···················· | ···················· | ···················· | |
| G | 7434 hCoV-19/USA/FL-BPHL-0919/2020\|EPI_ISL_517809\|2020- | ···················· | ···················· | ···················· | ···················· | ···················· | ···················· | |
| G | 7436 hCoV-19/USA/FL-BPHL-0922/2020\|EPI_ISL_517812\|2020- | ···················· | ···················· | ···················· | ···················· | NNN················· | ···················· | 1 |
| G | 7437 hCoV-19/USA/FL-BPHL-0923/2020\|EPI_ISL_517813\|2020- | ···················· | ···················· | ···················· | ···················· | ···················· | ···················· | |
| G | 7438 hCoV-19/USA/FL-BPHL-0924/2020\|EPI_ISL_517814\|2020- | ···················· | ···················· | ···················· | ···················· | ···················· | ···················· | |
| G | 7439 hCoV-19/USA/FL-BPHL-0925/2020\|EPI_ISL_517815\|2020- | ···················· | ···················· | ···················· | ···················· | ···················· | ···················· | |
| G | 7440 hCoV-19/USA/FL-BPHL-0926/2020\|EPI_ISL_517816\|2020- | ···················· | ···················· | ···················· | ···················· | ···················· | ···················· | |

G 7441 hCoV-19/USA/FL-BPHL-0927/2020|EPI_ISL_517817|2020- .................... .................... .................... .................... .................... ....................
G 7444 hCoV-19/USA/FL-BPHL-0933/2020|EPI_ISL_517823|2020- .................... .................... .................... .................... .................... ....................
G 7445 hCoV-19/USA/FL-BPHL-0935/2020|EPI_ISL_517825|2020- .................... .................... .................... .................... .................... ....................
G 7446 hCoV-19/USA/FL-BPHL-0937/2020|EPI_ISL_517827|2020- .................... .................... .................... .................... .................... ....................
G 7447 hCoV-19/USA/FL-BPHL-0938/2020|EPI_ISL_517828|2020- .................... .................... .................... .................... .................... ....................
G 7450 hCoV-19/USA/FL-BPHL-0941/2020|EPI_ISL_517831|2020- .................... .................... .................... .................... .................... ....................
G 7451 hCoV-19/USA/FL-BPHL-0942/2020|EPI_ISL_517832|2020- .................... .................... .................... .................... .................... ....................
G 7452 hCoV-19/USA/FL-BPHL-0943/2020|EPI_ISL_517833|2020- .................... .................... .................... .................... .................... ....................
G 7454 hCoV-19/USA/FL-BPHL-0945/2020|EPI_ISL_517835|2020- .................... .................... .................... .................... .................... ....................
G 7455 hCoV-19/USA/FL-BPHL-0946/2020|EPI_ISL_517836|2020- .................... .................... .................... .................... .................... ....................
G 7456 hCoV-19/USA/FL-BPHL-0992/2020|EPI_ISL_517882|2020- .................... .................... .................... .................... .................... ....................
G 7457 hCoV-19/USA/FL-BPHL-0993/2020|EPI_ISL_517883|2020- .................... .................... .................... .................... .................... ....................
G 7458 hCoV-19/USA/FL-BPHL-1000/2020|EPI_ISL_517890|2020- .................... .................... .................... .................... .................... ....................
G 7462 hCoV-19/USA/FL-BPHL-1005/2020|EPI_ISL_517895|2020- .................... .................... .................... .................... .................... ....................
G 7463 hCoV-19/USA/FL-BPHL-1006/2020|EPI_ISL_517896|2020- .................... .................... .................... .................... .................... ....................
G 7464 hCoV-19/USA/FL-BPHL-1007/2020|EPI_ISL_517897|2020- .................... .................... .................... .................... .................... ....................
G 7465 hCoV-19/USA/FL-BPHL-1009/2020|EPI_ISL_517899|2020- .................... .................... .................... .................... .................... ....................
G 7466 hCoV-19/Australia/VIC3338/2020|EPI_ISL_521017|2020 .................... .................... .................... .................... .................... ....................
G 7467 hCoV-19/Australia/VIC3339/2020|EPI_ISL_521018|2020 .................... .................... .................... .................... .................... ....................
G 7468 hCoV-19/Australia/VIC3340/2020|EPI_ISL_521019|2020 .................... .................... .................... .................... .................... ....................
G 7469 hCoV-19/Australia/VIC3341/2020|EPI_ISL_521020|2020 .................... .................... .................... .................... .................... ....................
G 7470 hCoV-19/Australia/VIC3342/2020|EPI_ISL_521021|2020 .................... .................... .................... .................... .................... ....................
G 7471 hCoV-19/Australia/VIC3343/2020|EPI_ISL_521022|2020 .................... .................... .................... .................... .................... ....................
G 7472 hCoV-19/Australia/VIC3344/2020|EPI_ISL_521023|2020 .................... .................... .................... .................... .................... ....................
G 7473 hCoV-19/Australia/VIC3345/2020|EPI_ISL_521024|2020 .................... .................... .................... .................... .................... ....................
G 7474 hCoV-19/Australia/VIC3346/2020|EPI_ISL_521025|2020 .................... .................... .................... .................... .................... ....................
G 7475 hCoV-19/Australia/VIC3347/2020|EPI_ISL_521026|2020 .................... .................... .................... .................... .................... ....................
G 7476 hCoV-19/Australia/VIC3348/2020|EPI_ISL_521027|2020 .................... .................... .................... .................... .................... ....................
G 7477 hCoV-19/Australia/VIC3349/2020|EPI_ISL_521028|2020 .................... .................... .................... .................... .................... ....................
G 7478 hCoV-19/Australia/VIC3350/2020|EPI_ISL_521029|2020 .................... .................... .................... .................... .................... ....................
G 7479 hCoV-19/Australia/VIC3351/2020|EPI_ISL_521030|2020 .................... .................... .................... .................... .................... ....................
G 7480 hCoV-19/Australia/VIC3352/2020|EPI_ISL_521031|2020 .................... .................... .................... .................... .................... ....................
G 7481 hCoV-19/Australia/VIC3353/2020|EPI_ISL_521032|2020 .................... .................... .................... .................... .................... ....................
G 7482 hCoV-19/Australia/VIC3354/2020|EPI_ISL_521033|2020 .................... .................... .................... .................... .................... ....................
G 7483 hCoV-19/Australia/VIC3355/2020|EPI_ISL_521034|2020 .................... .................... .................... .................... .................... ....................
G 7484 hCoV-19/Australia/VIC3356/2020|EPI_ISL_521035|2020 .................... .................... .................... .................... .................... ....................
G 7485 hCoV-19/Australia/VIC3357/2020|EPI_ISL_521036|2020 .................... .................... .................... .................... .................... ....................
G 7486 hCoV-19/Australia/VIC3358/2020|EPI_ISL_521037|2020 .................... .................... .................... .................... .................... ....................
G 7487 hCoV-19/Australia/VIC3359/2020|EPI_ISL_521038|2020 .................... .................... .................... .................... .................... ....................
G 7488 hCoV-19/Australia/VIC3360/2020|EPI_ISL_521039|2020 .................... .................... .................... .................... .................... ....................
G 7489 hCoV-19/Australia/VIC3361/2020|EPI_ISL_521040|2020 .................... .................... .................... .................... .................... ....................
G 7490 hCoV-19/Australia/VIC3362/2020|EPI_ISL_521041|2020 .................... .................... .................... .................... .................... ....................
G 7491 hCoV-19/Australia/VIC3363/2020|EPI_ISL_521042|2020 .................... .................... .................... .................... .................... ....................
G 7492 hCoV-19/Australia/VIC3364/2020|EPI_ISL_521043|2020 .................... .................... .................... .................... .................... ....................
G 7493 hCoV-19/Australia/VIC3365/2020|EPI_ISL_521044|2020 .................... .................... .................... .................... .................... ....................
G 7494 hCoV-19/Australia/VIC3366/2020|EPI_ISL_521045|2020 .................... .................... .................... .................... .................... ....................
G 7496 hCoV-19/Australia/VIC3368/2020|EPI_ISL_521047|2020 .................... .................... .................... .................... .................... ....................
G 7497 hCoV-19/Australia/VIC3369/2020|EPI_ISL_521048|2020 .................... .................... .................... .................... .................... ....................
G 7498 hCoV-19/Australia/VIC3370/2020|EPI_ISL_521049|2020 .................... .................... .................... .................... .................... ....................
G 7499 hCoV-19/Australia/VIC3371/2020|EPI_ISL_521050|2020 .................... .................... .................... .................... .................... ....................
G 7500 hCoV-19/Australia/VIC3372/2020|EPI_ISL_521051|2020 .................... .................... .................... .................... .................... ....................
G 7501 hCoV-19/Australia/VIC3373/2020|EPI_ISL_521052|2020 .................... .................... .................... .................... .................... ....................
G 7502 hCoV-19/Australia/VIC3374/2020|EPI_ISL_521053|2020 .................... .................... .................... .................... .................... ....................
G 7503 hCoV-19/Australia/VIC3375/2020|EPI_ISL_521054|2020 .................... .................... .................... .................... .................... ....................
G 7504 hCoV-19/Australia/VIC3376/2020|EPI_ISL_521055|2020 .................... .................... .................... .................... .................... ....................
G 7505 hCoV-19/Australia/VIC3377/2020|EPI_ISL_521056|2020 .................... .................... .................... .................... .................... ....................
G 7506 hCoV-19/Australia/VIC3378/2020|EPI_ISL_521057|2020 .................... .................... .................... .................... .................... ....................
G 7507 hCoV-19/Australia/VIC3379/2020|EPI_ISL_521058|2020 .................... .................... .................... .................... .................... ....................
G 7508 hCoV-19/Australia/VIC3380/2020|EPI_ISL_521059|2020 .................... .................... .................... .................... .................... ....................
G 7509 hCoV-19/Australia/VIC3381/2020|EPI_ISL_521060|2020 .................... .................... .................... .................... .................... ....................
G 7510 hCoV-19/Australia/VIC3382/2020|EPI_ISL_521061|2020 .................... .................... .................... .................... .................... ....................
G 7511 hCoV-19/Australia/VIC3383/2020|EPI_ISL_521062|2020 .................... .................... .................... .................... .................... ....................
G 7512 hCoV-19/Australia/VIC3384/2020|EPI_ISL_521063|2020 .................... .................... .................... .................... .................... ....................
G 7513 hCoV-19/Australia/VIC3385/2020|EPI_ISL_521064|2020 .................... .................... .................... .................... .................... ....................
G 7514 hCoV-19/Australia/VIC3386/2020|EPI_ISL_521065|2020 .................... .................... .................... .................... .................... ....................
G 7515 hCoV-19/Australia/VIC3387/2020|EPI_ISL_521066|2020 .................... .................... .................... .................... .................... ....................
G 7516 hCoV-19/Australia/VIC3388/2020|EPI_ISL_521067|2020 .................... .................... .................... .................... .................... ....................
G 7517 hCoV-19/Australia/VIC3389/2020|EPI_ISL_521068|2020 .................... .................... .................... .................... .................... ....................
G 7518 hCoV-19/Australia/VIC3390/2020|EPI_ISL_521069|2020 .................... .................... .................... .................... .................... ....................
G 7519 hCoV-19/Australia/VIC3391/2020|EPI_ISL_521070|2020 .................... .................... .................... .................... .................... ....................
G 7520 hCoV-19/Australia/VIC3392/2020|EPI_ISL_521071|2020 .................... .................... .................... .................... .................... ....................
G 7521 hCoV-19/Australia/VIC3393/2020|EPI_ISL_521072|2020 .................... .................... .................... .................... .................... ....................
G 7522 hCoV-19/Australia/VIC3394/2020|EPI_ISL_521073|2020 .................... .................... .................... .................... .................... ....................
G 7523 hCoV-19/Australia/VIC3395/2020|EPI_ISL_521074|2020 .................... .................... .................... .................... .................... ....................
G 7524 hCoV-19/Australia/VIC3397/2020|EPI_ISL_521076|2020 .................... .................... .................... .................... .................... ....................
G 7525 hCoV-19/Australia/VIC3398/2020|EPI_ISL_521077|2020 .................... .................... .................... .................... .................... ....................
G 7526 hCoV-19/Australia/VIC3399/2020|EPI_ISL_521078|2020 .................... .................... .................... .................... .................... ....................
G 7527 hCoV-19/Australia/VIC3400/2020|EPI_ISL_521079|2020 .................... .................... .................... .................... .................... ....................
G 7528 hCoV-19/Australia/VIC3401/2020|EPI_ISL_521080|2020 .................... .................... .................... .................... .................... ....................
G 7529 hCoV-19/Australia/VIC3402/2020|EPI_ISL_521081|2020 .................... .................... .................... .................... .................... ....................

| | | | | | | | | |
|---|---|---|---|---|---|---|---|---|
| G | 7530 hCoV-19/Australia/VIC3403/2020\|EPI_ISL_521082\|2020 | ···················· | ···················· | ···················· | ···················· | ···················· | ···················· | |
| G | 7531 hCoV-19/Australia/VIC3404/2020\|EPI_ISL_521083\|2020 | ···················· | ···················· | ···················· | ···················· | ···················· | ···················· | |
| G | 7532 hCoV-19/Australia/VIC3405/2020\|EPI_ISL_521084\|2020 | ···················· | ···················· | ···················· | ···················· | ···················· | ···················· | |
| G | 7533 hCoV-19/Australia/VIC3406/2020\|EPI_ISL_521085\|2020 | ···················· | ···················· | ···················· | ···················· | ···················· | ···················· | |
| G | 7534 hCoV-19/Australia/VIC3407/2020\|EPI_ISL_521086\|2020 | ···················· | ···················· | ···················· | ···················· | ···················· | ···················· | |
| G | 7535 hCoV-19/Australia/VIC3409/2020\|EPI_ISL_521087\|2020 | ···················· | ···················· | ···················· | ···················· | ···················· | ···················· | |
| G | 7536 hCoV-19/Australia/VIC3411/2020\|EPI_ISL_521088\|2020 | ···················· | ···················· | ···················· | ···················· | ···················· | ···················· | |
| G | 7538 hCoV-19/Australia/VIC3417/2020\|EPI_ISL_521091\|2020 | ···················· | ···················· | ···················· | ···················· | ···················· | ···················· | |
| G | 7540 hCoV-19/Australia/VIC3419/2020\|EPI_ISL_521093\|2020 | ···················· | ···················· | ···················· | ···················· | ···················· | ···················· | |
| G | 7541 hCoV-19/Australia/VIC3420/2020\|EPI_ISL_521094\|2020 | ···················· | ···················· | ···················· | ···················· | NNN················· | ···················· | 1 |
| G | 7543 hCoV-19/Australia/VIC3425/2020\|EPI_ISL_521096\|2020 | ···················· | ···················· | ···················· | ···················· | ···················· | ···················· | |
| G | 7546 hCoV-19/Australia/VIC3429/2020\|EPI_ISL_521099\|2020 | ···················· | ···················· | ···················· | ···················· | ···················· | ···················· | |
| G | 7547 hCoV-19/USA/UT-03028/2020\|EPI_ISL_522300\|2020-05-0 | ···················· | ···················· | ···················· | ···················· | ···················· | ···················· | |
| G | 7548 hCoV-19/USA/UT-03030/2020\|EPI_ISL_522302\|2020-05-0 | ···················· | ···················· | ···················· | ···················· | ···················· | ···················· | |
| G | 7549 hCoV-19/USA/UT-03031/2020\|EPI_ISL_522303\|2020-05-0 | ···················· | ···················· | ···················· | ···················· | ···················· | ···················· | |
| G | 7550 hCoV-19/USA/UT-03032/2020\|EPI_ISL_522304\|2020-05-0 | ···················· | ···················· | ···················· | ···················· | ···················· | ···················· | |
| G | 7551 hCoV-19/USA/UT-03033/2020\|EPI_ISL_522305\|2020-05-0 | ···················· | ···················· | ···················· | ···················· | ···················· | ···················· | |
| G | 7552 hCoV-19/USA/UT-03035/2020\|EPI_ISL_522307\|2020-05-0 | ···················· | ···················· | ···················· | ···················· | ···················· | ···················· | |
| G | 7553 hCoV-19/USA/UT-03039/2020\|EPI_ISL_522311\|2020-05-0 | ···················· | ···················· | ···················· | ···················· | ···················· | ···················· | |
| G | 7554 hCoV-19/USA/UT-03041/2020\|EPI_ISL_522312\|2020-05-0 | ···················· | ···················· | ···················· | ···················· | ···················· | ···················· | |
| G | 7555 hCoV-19/USA/UT-03044/2020\|EPI_ISL_522315\|2020-05-0 | ···················· | ···················· | ···················· | ···················· | ···················· | ···················· | |
| G | 7556 hCoV-19/USA/UT-04225/2020\|EPI_ISL_522322\|2020-05-1 | ···················· | ···················· | ···················· | ···················· | ···················· | ···················· | |
| G | 7557 hCoV-19/USA/UT-04226/2020\|EPI_ISL_522323\|2020-05-1 | ···················· | ···················· | ···················· | ···················· | ···················· | ···················· | |
| G | 7558 hCoV-19/Belgium/rega-0309752/2020\|EPI_ISL_522349\|2 | ···················· | ···················· | ···················· | ···················· | ···················· | ···················· | |
| G | 7559 hCoV-19/Belgium/rega-0710751/2020\|EPI_ISL_522350\|2 | ···················· | ···················· | ···················· | ···················· | ···················· | ···················· | |
| G | 7560 hCoV-19/USA/TX-DSHS-0094/2020\|EPI_ISL_522378\|2020- | ···················· | ···················· | ···················· | ···················· | ···················· | ···················· | |
| G | 7561 hCoV-19/USA/TX-DSHS-0110/2020\|EPI_ISL_522393\|2020- | ···················· | ···················· | ···················· | ···················· | ···················· | ···················· | |
| G | 7562 hCoV-19/USA/TX-DSHS-0111/2020\|EPI_ISL_522394\|2020- | ···················· | ···················· | ···················· | ···················· | ···················· | ···················· | |
| G | 7563 hCoV-19/USA/AK-PHL346/2020\|EPI_ISL_522396\|2020-05- | ···················· | ···················· | ···················· | ···················· | ···················· | ···················· | |
| G | 7564 hCoV-19/USA/AK-PHL403/2020\|EPI_ISL_522397\|2020-06- | ···················· | ···················· | ···················· | ···················· | ···················· | ···················· | |
| G | 7565 hCoV-19/USA/AK-PHL414/2020\|EPI_ISL_522398\|2020-06- | ···················· | ···················· | ···················· | ···················· | ···················· | ···················· | |
| G | 7566 hCoV-19/USA/AK-PHL442/2020\|EPI_ISL_522399\|2020-06- | ···················· | ···················· | ···················· | ···················· | ···················· | ···················· | |
| G | 7567 hCoV-19/Netherlands/ZH-EMC-258/2020\|EPI_ISL_523600 | ···················· | ···················· | ···················· | ···················· | ···················· | ···················· | |
| G | 7568 hCoV-19/Netherlands/ZH-EMC-259/2020\|EPI_ISL_523601 | ···················· | ···················· | ···················· | ···················· | ···················· | ···················· | |
| G | 7569 hCoV-19/Netherlands/ZH-EMC-260/2020\|EPI_ISL_523602 | ···················· | ···················· | ···················· | ···················· | ···················· | ···················· | |
| G | 7570 hCoV-19/Netherlands/ZH-EMC-261/2020\|EPI_ISL_523603 | ···················· | ···················· | ···················· | ···················· | ···················· | ···················· | |
| G | 7571 hCoV-19/Netherlands/ZH-EMC-262/2020\|EPI_ISL_523604 | ···················· | ···················· | ···················· | ···················· | ···················· | ···················· | |
| G | 7572 hCoV-19/Netherlands/ZH-EMC-263/2020\|EPI_ISL_523605 | ···················· | ···················· | ···················· | ···················· | ···················· | ···················· | |
| G | 7573 hCoV-19/Netherlands/ZH-EMC-264/2020\|EPI_ISL_523606 | ···················· | ···················· | ···················· | ···················· | ···················· | ···················· | |
| G | 7574 hCoV-19/Netherlands/ZH-EMC-265/2020\|EPI_ISL_523607 | ···················· | ···················· | ···················· | ···················· | ···················· | ···················· | |
| G | 7575 hCoV-19/Netherlands/ZH-EMC-266/2020\|EPI_ISL_523608 | ···················· | ···················· | ···················· | ···················· | ···················· | ···················· | |
| G | 7576 hCoV-19/Netherlands/ZH-EMC-267/2020\|EPI_ISL_523609 | ···················· | ···················· | ···················· | ···················· | ···················· | ···················· | |
| G | 7577 hCoV-19/Netherlands/ZH-EMC-268/2020\|EPI_ISL_523610 | ···················· | ···················· | ···················· | ···················· | ···················· | ···················· | |
| G | 7578 hCoV-19/Netherlands/ZH-EMC-269/2020\|EPI_ISL_523611 | ···················· | ···················· | ···················· | ···················· | ···················· | ···················· | |
| G | 7579 hCoV-19/Netherlands/ZH-EMC-270/2020\|EPI_ISL_523612 | ···················· | ···················· | ···················· | ···················· | ···················· | ···················· | |
| G | 7580 hCoV-19/Netherlands/ZH-EMC-271/2020\|EPI_ISL_523613 | ···················· | ···················· | ···················· | ···················· | ···················· | ···················· | |
| G | 7581 hCoV-19/Netherlands/ZH-EMC-272/2020\|EPI_ISL_523614 | ···················· | ···················· | ···················· | ···················· | ···················· | ···················· | |
| G | 7582 hCoV-19/Netherlands/ZH-EMC-274/2020\|EPI_ISL_523616 | ···················· | ···················· | ···················· | ···················· | ···················· | ···················· | |
| G | 7583 hCoV-19/Netherlands/ZH-EMC-276/2020\|EPI_ISL_523618 | ···················· | ···················· | ···················· | ···················· | ···················· | ···················· | |
| G | 7584 hCoV-19/Netherlands/ZH-EMC-277/2020\|EPI_ISL_523619 | ···················· | ···················· | ···················· | ···················· | ···················· | ···················· | |
| G | 7585 hCoV-19/Netherlands/ZH-EMC-278/2020\|EPI_ISL_523620 | ···················· | ···················· | ···················· | ···················· | ···················· | ···················· | |
| G | 7586 hCoV-19/Netherlands/ZH-EMC-279/2020\|EPI_ISL_523621 | ···················· | ···················· | ···················· | ···················· | ···················· | ···················· | |
| G | 7587 hCoV-19/Netherlands/ZH-EMC-280/2020\|EPI_ISL_523622 | ···················· | ···················· | ···················· | ···················· | ···················· | ···················· | |
| G | 7588 hCoV-19/Netherlands/ZH-EMC-281/2020\|EPI_ISL_523623 | ···················· | ···················· | ···················· | ···················· | ···················· | ···················· | |
| G | 7589 hCoV-19/Netherlands/ZH-EMC-283/2020\|EPI_ISL_523625 | ···················· | ···················· | ···················· | ···················· | ···················· | ···················· | |
| G | 7590 hCoV-19/Netherlands/ZH-EMC-284/2020\|EPI_ISL_523626 | ···················· | ···················· | ···················· | ···················· | ···················· | ···················· | |
| G | 7591 hCoV-19/Netherlands/ZH-EMC-285/2020\|EPI_ISL_523627 | ···················· | ···················· | ···················· | ···················· | ···················· | ···················· | |
| G | 7592 hCoV-19/Netherlands/ZH-EMC-286/2020\|EPI_ISL_523628 | ···················· | ···················· | ···················· | ···················· | ···················· | ···················· | |
| G | 7593 hCoV-19/Netherlands/ZH-EMC-287/2020\|EPI_ISL_523629 | ···················· | ···················· | ···················· | ···················· | ···················· | ···················· | |
| G | 7594 hCoV-19/Netherlands/ZH-EMC-288/2020\|EPI_ISL_523630 | ···················· | ···················· | ···················· | ···················· | ···················· | ···················· | |
| G | 7595 hCoV-19/Netherlands/ZH-EMC-289/2020\|EPI_ISL_523631 | ···················· | ···················· | ···················· | ···················· | ···················· | ···················· | |
| G | 7596 hCoV-19/Netherlands/ZH-EMC-290/2020\|EPI_ISL_523632 | ···················· | ···················· | ···················· | ···················· | ···················· | ···················· | |
| G | 7597 hCoV-19/Netherlands/ZH-EMC-291/2020\|EPI_ISL_523633 | ···················· | ···················· | ···················· | ···················· | ···················· | ···················· | |
| G | 7598 hCoV-19/Netherlands/ZH-EMC-292/2020\|EPI_ISL_523634 | ···················· | ···················· | ···················· | ···················· | ···················· | ···················· | |
| G | 7599 hCoV-19/Netherlands/ZH-EMC-293/2020\|EPI_ISL_523635 | ···················· | ···················· | ···················· | ···················· | ···················· | ···················· | |
| G | 7600 hCoV-19/Netherlands/ZH-EMC-294/2020\|EPI_ISL_523636 | ···················· | ···················· | ···················· | ···················· | ···················· | ···················· | |
| G | 7601 hCoV-19/Netherlands/ZH-EMC-295/2020\|EPI_ISL_523637 | ···················· | ···················· | ···················· | ···················· | ···················· | ···················· | |
| G | 7602 hCoV-19/Netherlands/ZH-EMC-296/2020\|EPI_ISL_523638 | ···················· | ···················· | ···················· | ···················· | ···················· | ···················· | |
| G | 7603 hCoV-19/Netherlands/ZH-EMC-297/2020\|EPI_ISL_523639 | ···················· | ···················· | ···················· | ···················· | ···················· | ···················· | |
| G | 7604 hCoV-19/Netherlands/ZH-EMC-298/2020\|EPI_ISL_523640 | ···················· | ···················· | ···················· | ···················· | ···················· | ···················· | |
| G | 7605 hCoV-19/Netherlands/ZH-EMC-299/2020\|EPI_ISL_523641 | ···················· | ···················· | ···················· | ···················· | ···················· | ···················· | |
| G | 7606 hCoV-19/Netherlands/ZH-EMC-301/2020\|EPI_ISL_523643 | ···················· | ···················· | ···················· | ···················· | ···················· | ···················· | |
| G | 7607 hCoV-19/Netherlands/ZH-EMC-302/2020\|EPI_ISL_523644 | ···················· | ···················· | ···················· | ···················· | ···················· | ···················· | |
| G | 7608 hCoV-19/Netherlands/ZH-EMC-303/2020\|EPI_ISL_523645 | ···················· | ···················· | ···················· | ···················· | ···················· | ···················· | |
| G | 7609 hCoV-19/Netherlands/ZH-EMC-304/2020\|EPI_ISL_523646 | ···················· | ···················· | ···················· | ···················· | ···················· | ···················· | |
| G | 7610 hCoV-19/Netherlands/ZH-EMC-305/2020\|EPI_ISL_523647 | ···················· | ···················· | ···················· | ···················· | ···················· | ···················· | |
| G | 7611 hCoV-19/Netherlands/ZH-EMC-306/2020\|EPI_ISL_523648 | ···················· | ···················· | ···················· | ···················· | ···················· | ···················· | |
| G | 7612 hCoV-19/Netherlands/ZH-EMC-307/2020\|EPI_ISL_523649 | ···················· | ···················· | ···················· | ···················· | ···················· | ···················· | |
| G | 7613 hCoV-19/Netherlands/ZH-EMC-338/2020\|EPI_ISL_523679 | ···················· | ···················· | ···················· | ···················· | ···················· | ···················· | |
| G | 7614 hCoV-19/Netherlands/ZH-EMC-339/2020\|EPI_ISL_523680 | ···················· | ···················· | ···················· | ···················· | ···················· | ···················· | |

| | | | | | | | | |
|---|---|---|---|---|---|---|---|---|
| G | 7615 | hCoV-19/Netherlands/ZH-EMC-340/2020\|EPI_ISL_523681 | .................... | .................... | .................... | .................... | .................... | .................... |
| G | 7616 | hCoV-19/Netherlands/ZH-EMC-341/2020\|EPI_ISL_523682 | .................... | .................... | .................... | .................... | .................... | .................... |
| G | 7617 | hCoV-19/Netherlands/ZH-EMC-342/2020\|EPI_ISL_523683 | .................... | .................... | .................... | .................... | .................... | .................... |
| G | 7618 | hCoV-19/Netherlands/ZH-EMC-343/2020\|EPI_ISL_523684 | .................... | .................... | .................... | .................... | .................... | .................... |
| G | 7619 | hCoV-19/Netherlands/ZH-EMC-344/2020\|EPI_ISL_523685 | .................... | .................... | .................... | .................... | .................... | .................... |
| G | 7620 | hCoV-19/Netherlands/ZH-EMC-345/2020\|EPI_ISL_523686 | .................... | .................... | .................... | .................... | .................... | .................... |
| G | 7621 | hCoV-19/Netherlands/ZH-EMC-346/2020\|EPI_ISL_523687 | .................... | .................... | .................... | .................... | .................... | .................... |
| G | 7622 | hCoV-19/Netherlands/ZH-EMC-348/2020\|EPI_ISL_523689 | .................... | .................... | .................... | .................... | .................... | .................... |
| G | 7623 | hCoV-19/Netherlands/ZH-EMC-349/2020\|EPI_ISL_523690 | .................... | .................... | .................... | .................... | .................... | .................... |
| G | 7624 | hCoV-19/Netherlands/ZH-EMC-351/2020\|EPI_ISL_523692 | .................... | .................... | .................... | .................... | .................... | .................... |
| G | 7625 | hCoV-19/Netherlands/ZH-EMC-353/2020\|EPI_ISL_523693 | .................... | .................... | .................... | .................... | .................... | .................... |
| G | 7626 | hCoV-19/Netherlands/ZH-EMC-354/2020\|EPI_ISL_523694 | .................... | .................... | .................... | .................... | .................... | .................... |
| G | 7627 | hCoV-19/Netherlands/ZH-EMC-355/2020\|EPI_ISL_523695 | .................... | .................... | .................... | .................... | .................... | .................... |
| G | 7628 | hCoV-19/Netherlands/ZH-EMC-356/2020\|EPI_ISL_523696 | .................... | .................... | .................... | .................... | .................... | .................... |
| G | 7629 | hCoV-19/Netherlands/ZH-EMC-357/2020\|EPI_ISL_523697 | .................... | .................... | .................... | .................... | .................... | .................... |
| G | 7630 | hCoV-19/Netherlands/ZH-EMC-358/2020\|EPI_ISL_523698 | .................... | .................... | .................... | .................... | .................... | .................... |
| G | 7631 | hCoV-19/Netherlands/ZH-EMC-359/2020\|EPI_ISL_523699 | .................... | .................... | .................... | .................... | .................... | .................... |
| G | 7632 | hCoV-19/USA/UT-UPHL-2009318/2020\|EPI_ISL_524923\|20 | .................... | .................... | .................... | .................... | .................... | .................... |
| G | 7633 | hCoV-19/USA/UT-UPHL-2009324/2020\|EPI_ISL_524925\|20 | .................... | .................... | .................... | .................... | .................... | .................... |
| G | 7634 | hCoV-19/USA/UT-UPHL-2009325/2020\|EPI_ISL_524926\|20 | .................... | .................... | .................... | .................... | .................... | .................... |
| G | 7635 | hCoV-19/USA/UT-UPHL-2009326/2020\|EPI_ISL_524927\|20 | .................... | .................... | .................... | .................... | .................... | .................... |
| G | 7636 | hCoV-19/USA/UT-UPHL-2009327/2020\|EPI_ISL_524928\|20 | .................... | .................... | .................... | .................... | .................... | .................... |
| G | 7637 | hCoV-19/USA/UT-UPHL-2009330/2020\|EPI_ISL_524931\|20 | .................... | .................... | .................... | .................... | .................... | .................... |
| G | 7638 | hCoV-19/USA/UT-UPHL-2009331/2020\|EPI_ISL_524932\|20 | .................... | .................... | .................... | .................... | .........A.......... | .................... |
| G | 7639 | hCoV-19/USA/UT-UPHL-2009333/2020\|EPI_ISL_524933\|20 | .................... | .................... | .................... | .................... | .................... | .................... |
| G | 7640 | hCoV-19/USA/UT-UPHL-2009334/2020\|EPI_ISL_524934\|20 | .................... | .................... | .................... | .................... | .........A.......... | .................... |
| G | 7641 | hCoV-19/USA/UT-UPHL-2009335/2020\|EPI_ISL_524935\|20 | .................... | .................... | .................... | .................... | .................... | .................... |
| G | 7642 | hCoV-19/USA/UT-UPHL-2009338/2020\|EPI_ISL_524938\|20 | .................... | .................... | .................... | .................... | .................... | .................... |
| G | 7643 | hCoV-19/USA/UT-UPHL-2009341/2020\|EPI_ISL_524939\|20 | .................... | .................... | .................... | .................... | .................... | .................... |
| G | 7644 | hCoV-19/USA/UT-UPHL-2009342/2020\|EPI_ISL_524940\|20 | .................... | .................... | .................... | .................... | .................... | .................... |
| G | 7645 | hCoV-19/USA/UT-UPHL-2009344/2020\|EPI_ISL_524942\|20 | .................... | .................... | .................... | .................... | .................... | .................... |
| G | 7646 | hCoV-19/USA/UT-UPHL-2009345/2020\|EPI_ISL_524943\|20 | .................... | .................... | .................... | .................... | .................... | .................... |
| G | 7647 | hCoV-19/USA/UT-UPHL-2009346/2020\|EPI_ISL_524944\|20 | .................... | .................... | .................... | .................... | .................... | .................... |
| G | 7648 | hCoV-19/USA/UT-UPHL-2009347/2020\|EPI_ISL_524945\|20 | .................... | .................... | .................... | .................... | .................... | .................... |
| G | 7649 | hCoV-19/USA/UT-UPHL-2009349/2020\|EPI_ISL_524946\|20 | .................... | .................... | .................... | .................... | .................... | .................... |
| G | 7650 | hCoV-19/USA/UT-UPHL-2009351/2020\|EPI_ISL_524948\|20 | .................... | .................... | .................... | .................... | .................... | .................... |
| G | 7651 | hCoV-19/USA/UT-UPHL-2009352/2020\|EPI_ISL_524949\|20 | .................... | .................... | .................... | .................... | .................... | .................... |
| G | 7652 | hCoV-19/USA/UT-UPHL-2009355/2020\|EPI_ISL_524950\|20 | .................... | .................... | .................... | .................... | .................... | .................... |
| G | 7653 | hCoV-19/USA/UT-UPHL-2009357/2020\|EPI_ISL_524952\|20 | .................... | .................... | .................... | .................... | .................... | .................... |
| G | 7654 | hCoV-19/USA/UT-UPHL-2009359/2020\|EPI_ISL_524954\|20 | .................... | .................... | .................... | .................... | .................... | .................... |
| G | 7655 | hCoV-19/USA/UT-UPHL-2009360/2020\|EPI_ISL_524955\|20 | .................... | .................... | .................... | .................... | .................... | .................... |
| G | 7656 | hCoV-19/USA/UT-UPHL-2009363/2020\|EPI_ISL_524957\|20 | .................... | .................... | .................... | .................... | .................... | .................... |
| G | 7657 | hCoV-19/USA/UT-UPHL-2009365/2020\|EPI_ISL_524958\|20 | .................... | .................... | .................... | .................... | .................... | .................... |
| G | 7658 | hCoV-19/USA/UT-UPHL-2009366/2020\|EPI_ISL_524959\|20 | .................... | .................... | .................... | .................... | .................... | .................... |
| G | 7659 | hCoV-19/USA/UT-UPHL-2009367/2020\|EPI_ISL_524960\|20 | .................... | .................... | .................... | .................... | .................... | .................... |
| G | 7660 | hCoV-19/USA/UT-UPHL-2009370/2020\|EPI_ISL_524963\|20 | .................... | .................... | .................... | .................... | .................... | .................... |
| G | 7661 | hCoV-19/USA/UT-UPHL-2009371/2020\|EPI_ISL_524964\|20 | .................... | .................... | .................... | .................... | .................... | .................... |
| G | 7662 | hCoV-19/USA/UT-UPHL-2009372/2020\|EPI_ISL_524965\|20 | .................... | .................... | .................... | .................... | .................... | .................... |
| G | 7663 | hCoV-19/USA/UT-UPHL-2009375/2020\|EPI_ISL_524968\|20 | .................... | .................... | .................... | .................... | .................... | .................... |
| G | 7664 | hCoV-19/USA/UT-UPHL-2009376/2020\|EPI_ISL_524969\|20 | .................... | .................... | .................... | .................... | .................... | .................... |
| G | 7665 | hCoV-19/USA/UT-UPHL-2009379/2020\|EPI_ISL_524971\|20 | .................... | .................... | .................... | .................... | .................... | .................... |
| G | 7666 | hCoV-19/USA/UT-UPHL-2008858/2020\|EPI_ISL_524976\|20 | .................... | .................... | .................... | .................... | .................... | .................... |
| G | 7667 | hCoV-19/USA/UT-UPHL-2008861/2020\|EPI_ISL_524979\|20 | .................... | .................... | .................... | .................... | .................... | .................... |
| G | 7668 | hCoV-19/United Arab Emirates/H29/2020\|EPI_ISL_5285 | .................... | .................... | .................... | .................... | .................... | .................... |
| G | 7669 | hCoV-19/Brazil/RJ-LVM161-2/2020\|EPI_ISL_528539\|202 | .................... | .................... | .................... | .................... | .................... | .................... |
| G | 7670 | hCoV-19/India/TG-CDFD-C1132/2020\|EPI_ISL_528548\|20 | .................... | .................... | .................... | .................... | .................... | .................... |
| G | 7671 | hCoV-19/India/TG-CDFD-C1138/2020\|EPI_ISL_528549\|20 | .................... | .................... | .................... | .................... | .................... | .................... |
| G | 7672 | hCoV-19/India/TG-CDFD-C1325/2020\|EPI_ISL_528555\|20 | .................... | .................... | .................... | .................... | .................... | .................... |
| G | 7673 | hCoV-19/India/TG-CDFD-C1327/2020\|EPI_ISL_528556\|20 | .................... | .................... | .................... | .................... | .................... | .................... |
| G | 7674 | hCoV-19/India/TG-CDFD-C1391/2020\|EPI_ISL_528560\|20 | .................... | .................... | .................... | .................... | .................... | .................... |
| G | 7675 | hCoV-19/India/TG-CDFD-C1408/2020\|EPI_ISL_528562\|20 | .................... | .................... | .................... | .................... | .................... | .................... |
| G | 7676 | hCoV-19/South Africa/KRISP-K002806/2020\|EPI_ISL_52 | .................... | .................... | .................... | .................... | .................... | .................... |
| G | 7677 | hCoV-19/South Africa/KRISP-K002807/2020\|EPI_ISL_52 | .................... | .................... | .................... | .................... | .................... | .................... |
| G | 7678 | hCoV-19/South Africa/KRISP-K002808/2020\|EPI_ISL_52 | .................... | .................... | .................... | .................... | .................... | .................... |
| G | 7679 | hCoV-19/USA/MI-MDHHS-SC21756/2020\|EPI_ISL_529874\|2 | .................... | .................... | .................... | .................... | .................... | .................... |
| G | 7681 | hCoV-19/Peru/CAJ-INS-057/2020\|EPI_ISL_517713\|2020- | .................... | .................... | .................... | .................... | .................... | .................... |
| G | 7682 | hCoV-19/USA/FL-BPHL-0949/2020\|EPI_ISL_517839\|2020- | .................... | .................... | .................... | .................... | .................... | .................... |
| G | 7683 | hCoV-19/USA/FL-BPHL-0950/2020\|EPI_ISL_517840\|2020- | .................... | .................... | .................... | .................... | .................... | .................... |
| G | 7687 | hCoV-19/USA/FL-BPHL-0956/2020\|EPI_ISL_517846\|2020- | .................... | .................... | .................... | .................... | .................... | .................... |
| G | 7688 | hCoV-19/USA/FL-BPHL-0957/2020\|EPI_ISL_517847\|2020- | .................... | .................... | .................... | .................... | .................... | .................... |
| G | 7689 | hCoV-19/USA/FL-BPHL-0959/2020\|EPI_ISL_517849\|2020- | .................... | .................... | .................... | .................... | .................... | .................... |
| G | 7690 | hCoV-19/USA/FL-BPHL-0960/2020\|EPI_ISL_517850\|2020- | .................... | .................... | .................... | .................... | .................... | .................... |
| G | 7691 | hCoV-19/USA/FL-BPHL-0962/2020\|EPI_ISL_517852\|2020- | .................... | .................... | .................... | .................... | .................... | .................... |
| G | 7692 | hCoV-19/USA/FL-BPHL-0964/2020\|EPI_ISL_517854\|2020- | .................... | .................... | .................... | .................... | .................... | .................... |
| G | 7693 | hCoV-19/USA/FL-BPHL-0967/2020\|EPI_ISL_517857\|2020- | .................... | .................... | .................... | .................... | .................... | .................... |
| G | 7694 | hCoV-19/USA/FL-BPHL-0968/2020\|EPI_ISL_517858\|2020- | .................... | .................... | .................... | .................... | .................... | .................... |
| G | 7695 | hCoV-19/USA/FL-BPHL-0969/2020\|EPI_ISL_517859\|2020- | .................... | .................... | .................... | .................... | .................... | .................... |
| G | 7696 | hCoV-19/USA/FL-BPHL-0970/2020\|EPI_ISL_517860\|2020- | .................... | .................... | .................... | .................... | .................... | .................... |
| G | 7698 | hCoV-19/USA/FL-BPHL-0972/2020\|EPI_ISL_517862\|2020- | .................... | .................... | .................... | .................... | .................... | .................... |
| G | 7700 | hCoV-19/USA/FL-BPHL-0974/2020\|EPI_ISL_517864\|2020- | .................... | .................... | .................... | .................... | .................... | .................... |

G 7701 hCoV-19/USA/FL-BPHL-0975/2020|EPI_ISL_517865|2020- ···················· ···················· ····················· ···················· ···················· ····················
G 7703 hCoV-19/USA/FL-BPHL-0977/2020|EPI_ISL_517867|2020- ···················· ···················· ····················· ···················· ···················· ····················
G 7704 hCoV-19/USA/FL-BPHL-0978/2020|EPI_ISL_517868|2020- ···················· ···················· ····················· ···················· ···················· ····················
G 7705 hCoV-19/USA/FL-BPHL-0979/2020|EPI_ISL_517869|2020- ···················· ···················· ····················· ···················· ···················· ····················
G 7708 hCoV-19/USA/FL-BPHL-0983/2020|EPI_ISL_517873|2020- ···················· ···················· ····················· ···················· ··N················· ····················
G 7709 hCoV-19/USA/FL-BPHL-0985/2020|EPI_ISL_517875|2020- ···················· ···················· ····················· ···················· ···················· ····················
G 7711 hCoV-19/USA/FL-BPHL-0988/2020|EPI_ISL_517878|2020- ···················· ···················· ····················· ···················· ···················· ····················
G 7712 hCoV-19/USA/FL-BPHL-0989/2020|EPI_ISL_517879|2020- ···················· ···················· ····················· ···················· ···················· ····················
G 7713 hCoV-19/USA/FL-BPHL-0990/2020|EPI_ISL_517880|2020- ···················· ···················· ····················· ···················· ···················· ····················
G 7714 hCoV-19/USA/FL-BPHL-0995/2020|EPI_ISL_517885|2020- ···················· ···················· ····················· ···················· ···················· ····················
G 7715 hCoV-19/USA/FL-BPHL-0998/2020|EPI_ISL_517888|2020- ···················· ···················· ····················· ···················· ···················· ····················
G 7717 hCoV-19/Australia/VIC2505/2020|EPI_ISL_522200|2020 ···················· ···················· ····················· ···················· NN·················· ····················
G 7719 hCoV-19/Australia/VIC2507/2020|EPI_ISL_522202|2020 ···················· ···················· ····················· ···················· ···················· ····················
G 7720 hCoV-19/Australia/VIC2508/2020|EPI_ISL_522203|2020 ···················· ···················· ····················· ···················· ···················· ····················
G 7721 hCoV-19/Australia/VIC2509/2020|EPI_ISL_522204|2020 ···················· ···················· ····················· ···················· ···················· ····················
G 7722 hCoV-19/Australia/VIC2510/2020|EPI_ISL_522205|2020 ···················· ···················· ····················· ···················· ···················· ····················
G 7723 hCoV-19/Australia/VIC2511/2020|EPI_ISL_522206|2020 ···················· ···················· ····················· ···················· ··N················· ····················
G 7724 hCoV-19/Australia/VIC2512/2020|EPI_ISL_522207|2020 ···················· ···················· ····················· ···················· ···················· ····················
G 7725 hCoV-19/Australia/VIC2513/2020|EPI_ISL_522208|2020 ···················· ···················· ····················· ···················· ···················· ····················
G 7726 hCoV-19/Australia/VIC2514/2020|EPI_ISL_522209|2020 ···················· ···················· ····················· ···················· ···················· ····················
G 7727 hCoV-19/Australia/VIC2515/2020|EPI_ISL_522210|2020 ···················· ···················· ····················· ···················· ···················· ····················
G 7728 hCoV-19/Australia/VIC2516/2020|EPI_ISL_522211|2020 ···················· ···················· ····················· ···················· ···················· ····················
G 7729 hCoV-19/Australia/VIC2517/2020|EPI_ISL_522212|2020 ···················· ···················· ····················· ···················· ···················· ····················
G 7730 hCoV-19/Australia/VIC2518/2020|EPI_ISL_522213|2020 ···················· ···················· ····················· ···················· ···················· ····················
G 7731 hCoV-19/Australia/VIC2519/2020|EPI_ISL_522214|2020 ···················· ···················· ····················· ···················· ···················· ····················
G 7732 hCoV-19/Australia/VIC2520/2020|EPI_ISL_522215|2020 ···················· ···················· ····················· ···················· ···················· ····················
G 7734 hCoV-19/Australia/VIC2523/2020|EPI_ISL_522217|2020 ···················· ···················· ····················· ···················· ···················· ····················
G 7735 hCoV-19/Australia/VIC2524/2020|EPI_ISL_522218|2020 ···················· ···················· ····················· ···················· ···················· ····················
G 7736 hCoV-19/Australia/VIC2525/2020|EPI_ISL_522219|2020 ···················· ···················· ····················· ···················· ···················· ····················
G 7737 hCoV-19/Australia/VIC2527/2020|EPI_ISL_522220|2020 ···················· ···················· ····················· ···················· ···················· ····················
G 7739 hCoV-19/Australia/VIC2529/2020|EPI_ISL_522222|2020 ···················· ···················· ····················· ···················· ···················· ····················
G 7740 hCoV-19/Australia/VIC2530/2020|EPI_ISL_522223|2020 ···················· ···················· ····················· ···················· ···················· ····················
G 7741 hCoV-19/Australia/VIC2531/2020|EPI_ISL_522224|2020 ···················· ···················· ····················· ···················· ···················· ····················
G 7742 hCoV-19/Australia/VIC2532/2020|EPI_ISL_522225|2020 ···················· ···················· ····················· ···················· ···················· ····················
G 7743 hCoV-19/Australia/VIC2533/2020|EPI_ISL_522226|2020 ···················· ···················· ····················· ···················· ···················· ····················
G 7744 hCoV-19/Australia/VIC2554/2020|EPI_ISL_522240|2020 ···················· ···················· ····················· ···················· ···················· ····················
G 7745 hCoV-19/Australia/VIC2559/2020|EPI_ISL_522243|2020 ···················· ···················· ····················· ···················· ···················· ····················
G 7746 hCoV-19/Australia/VIC2560/2020|EPI_ISL_522244|2020 ···················· ···················· ····················· ···················· ···················· ····················
G 7747 hCoV-19/Australia/VIC2571/2020|EPI_ISL_522249|2020 ···················· ···················· ····················· ···················· ···················· ····················
G 7748 hCoV-19/Australia/VIC2572/2020|EPI_ISL_522250|2020 ···················· ···················· ····················· ···················· ···················· ····················
G 7750 hCoV-19/Australia/VIC2574/2020|EPI_ISL_522252|2020 ···················· ···················· ····················· ···················· ···················· ····················
G 7751 hCoV-19/Australia/VIC2575/2020|EPI_ISL_522253|2020 ···················· ···················· ····················· ···················· ···················· ····················
G 7752 hCoV-19/USA/UT-02568/2020|EPI_ISL_522254|2020-04-2 ···················· ···················· ····················· ···················· ···················· ····················
G 7753 hCoV-19/USA/UT-02570/2020|EPI_ISL_522255|2020-04-2 ···················· ···················· ····················· ···················· ···················· ····················
G 7754 hCoV-19/USA/UT-02587/2020|EPI_ISL_522258|2020-04-2 ···················· ···················· ····················· ···················· ···················· ····················
G 7755 hCoV-19/USA/UT-02594/2020|EPI_ISL_522260|2020-04-2 ···················· ···················· ····················· ···················· ···················· ····················
G 7756 hCoV-19/USA/UT-02595/2020|EPI_ISL_522261|2020-04-2 ···················· ···················· ····················· ···················· ···················· ····················
G 7757 hCoV-19/USA/UT-02597/2020|EPI_ISL_522262|2020-04-2 ···················· ···················· ····················· ···················· ···················· ····················
G 7758 hCoV-19/USA/UT-02693/2020|EPI_ISL_522264|2020-04-2 ···················· ···················· ····················· ···················· ···················· ····················
G 7759 hCoV-19/USA/UT-02705/2020|EPI_ISL_522270|2020-04-2 ···················· ···················· ····················· ···················· ···················· ····················
G 7760 hCoV-19/USA/UT-02713/2020|EPI_ISL_522272|2020-04-2 ···················· ···················· ····················· ···················· ···················· ····················
G 7761 hCoV-19/USA/UT-02753/2020|EPI_ISL_522274|2020-04-2 ···················· ···················· ····················· ···················· ···················· ····················
G 7762 hCoV-19/USA/UT-02760/2020|EPI_ISL_522276|2020-04-2 ···················· ···················· ····················· ···················· ···················· ····················
G 7763 hCoV-19/USA/UT-02961/2020|EPI_ISL_522279|2020-05-0 ···················· ···················· ····················· ···················· ···················· ····················
G 7764 hCoV-19/USA/UT-02965/2020|EPI_ISL_522281|2020-05-0 ···················· ···················· ····················· ···················· ···················· ····················
G 7765 hCoV-19/USA/UT-02967/2020|EPI_ISL_522282|2020-05-0 ···················· ···················· ····················· ···················· ···················· ····················
G 7766 hCoV-19/USA/UT-02974/2020|EPI_ISL_522285|2020-05-0 ···················· ···················· ····················· ···················· ···················· ····················
G 7767 hCoV-19/USA/UT-02978/2020|EPI_ISL_522287|2020-05-0 ···················· ···················· ····················· ···················· ···················· ····················
G 7768 hCoV-19/USA/UT-02979/2020|EPI_ISL_522288|2020-05-0 ···················· ···················· ····················· ···················· ···················· ····················
G 7769 hCoV-19/USA/UT-02981/2020|EPI_ISL_522290|2020-05-0 ···················· ···················· ····················· ···················· ···················· ····················
G 7770 hCoV-19/USA/UT-02983/2020|EPI_ISL_522291|2020-05-0 ···················· ···················· ····················· ···················· ···················· ····················
G 7771 hCoV-19/USA/UT-02994/2020|EPI_ISL_522294|2020-05-0 ···················· ···················· ····················· ···················· ···················· ····················
G 7772 hCoV-19/USA/UT-03012/2020|EPI_ISL_522295|2020-05-0 ···················· ···················· ····················· ···················· ···················· ····················
G 7773 hCoV-19/USA/UT-03013/2020|EPI_ISL_522296|2020-05-0 ···················· ···················· ····················· ···················· ···················· ····················
G 7775 hCoV-19/Netherlands/un-EMC-745/2020|EPI_ISL_523501 ···················· ···················· ····················· ···················· ···················· ····················
G 7776 hCoV-19/Netherlands/un-EMC-746/2020|EPI_ISL_523502 ···················· ···················· ····················· ···················· ···················· ····················
G 7777 hCoV-19/Netherlands/un-EMC-747/2020|EPI_ISL_523503 ···················· ···················· ····················· ···················· ···················· ····················
G 7778 hCoV-19/Netherlands/un-EMC-748/2020|EPI_ISL_523504 ···················· ···················· ····················· ···················· ···················· ····················
G 7779 hCoV-19/Netherlands/un-EMC-749/2020|EPI_ISL_523505 ···················· ···················· ····················· ···················· ···················· ····················
G 7780 hCoV-19/Netherlands/un-EMC-750/2020|EPI_ISL_523506 ···················· ···················· ····················· ···················· ···················· ····················
G 7781 hCoV-19/Netherlands/un-EMC-751/2020|EPI_ISL_523507 ···················· ···················· ····················· ···················· ···················· ····················
G 7782 hCoV-19/Netherlands/un-EMC-752/2020|EPI_ISL_523508 ···················· ···················· ····················· ···················· ···················· ····················
G 7783 hCoV-19/Netherlands/un-EMC-753/2020|EPI_ISL_523509 ···················· ···········A········ ····················· ···················· ···················· ····················
G 7784 hCoV-19/Netherlands/un-EMC-757/2020|EPI_ISL_523513 ···················· ···················· ····················· ···················· ···················· ····················
G 7785 hCoV-19/Netherlands/un-EMC-758/2020|EPI_ISL_523514 ···················· ···················· ····················· ···················· ···················· ····················
G 7786 hCoV-19/Netherlands/un-EMC-759/2020|EPI_ISL_523515 ···················· ···················· ····················· ···················· ···················· ····················
G 7787 hCoV-19/Netherlands/un-EMC-760/2020|EPI_ISL_523516 ···················· ······A············· ····················· ···················· ···················· ····················
G 7788 hCoV-19/Netherlands/un-EMC-761/2020|EPI_ISL_523517 ···················· ·A·················· ····················· ···················· ···················· ···A················
G 7789 hCoV-19/Netherlands/un-EMC-762/2020|EPI_ISL_523518 ···················· ···················· ····················· ···················· ···················· ····················
G 7790 hCoV-19/Netherlands/un-EMC-764/2020|EPI_ISL_523520 ···················· ···················· ····················· ···················· ···················· ····················

| | | | | | | | |
|---|---|---|---|---|---|---|---|
| G | 7791 hCoV-19/Netherlands/un-EMC-765/2020\|EPI_ISL_523521 | ···················· | ···················· | ···················· | ···················· | ···················· | ···················· |
| G | 7792 hCoV-19/Netherlands/un-EMC-767/2020\|EPI_ISL_523523 | ···················· | ···················· | ···················· | ···················· | ···················· | ···················· |
| G | 7793 hCoV-19/Netherlands/un-EMC-768/2020\|EPI_ISL_523524 | ···················· | ···················· | ···················· | ···················· | ···················· | ···················· |
| G | 7794 hCoV-19/Netherlands/un-EMC-769/2020\|EPI_ISL_523525 | ···················· | ···················· | ···················· | ···················· | ···················· | ···················· |
| G | 7795 hCoV-19/Netherlands/un-EMC-770/2020\|EPI_ISL_523526 | ···················· | ···················· | ···················· | ···················· | ···················· | ···················· |
| G | 7796 hCoV-19/Netherlands/un-EMC-771/2020\|EPI_ISL_523527 | ···················· | ···················· | ···················· | ···················· | ···················· | ···················· |
| G | 7797 hCoV-19/Netherlands/UT-EMC-51/2020\|EPI_ISL_523528\| | ···················· | ···················· | ···················· | ···················· | ···················· | ···················· |
| G | 7798 hCoV-19/Netherlands/UT-EMC-52/2020\|EPI_ISL_523529\| | ···················· | ···················· | ···················· | ···················· | ···················· | ···················· |
| G | 7799 hCoV-19/Netherlands/UT-EMC-53/2020\|EPI_ISL_523530\| | ···················· | ···················· | ···················· | ···················· | ···················· | ···················· |
| G | 7800 hCoV-19/Netherlands/UT-EMC-54/2020\|EPI_ISL_523531\| | ···················· | ···················· | ···················· | ···················· | ···················· | ···················· |
| G | 7801 hCoV-19/Netherlands/UT-EMC-55/2020\|EPI_ISL_523532\| | ···················· | ··········A········· | ···················· | ···················· | ···················· | ···················· |
| G | 7802 hCoV-19/Netherlands/UT-EMC-56/2020\|EPI_ISL_523533\| | ···················· | ···················· | ···················· | ···················· | ···················· | ···················· |
| G | 7803 hCoV-19/Netherlands/UT-EMC-57/2020\|EPI_ISL_523534\| | ···················· | ···················· | ···················· | ···················· | ···················· | ···················· |
| G | 7804 hCoV-19/Netherlands/UT-EMC-59/2020\|EPI_ISL_523536\| | ···················· | ···················· | ···················· | ···················· | ···················· | ···················· |
| G | 7805 hCoV-19/Netherlands/UT-EMC-60/2020\|EPI_ISL_523537\| | ···················· | ···················· | ···················· | ···················· | ···················· | ···················· |
| G | 7806 hCoV-19/Netherlands/UT-EMC-62/2020\|EPI_ISL_523539\| | ···················· | ···················· | ···················· | ···················· | ···················· | ···················· |
| G | 7807 hCoV-19/Netherlands/UT-EMC-63/2020\|EPI_ISL_523540\| | ···················· | ···················· | ···················· | ···················· | ···················· | ···················· |
| G | 7808 hCoV-19/Netherlands/UT-EMC-64/2020\|EPI_ISL_523541\| | ···················· | ···················· | ···················· | ···················· | ···················· | ···················· |
| G | 7809 hCoV-19/Netherlands/UT-EMC-65/2020\|EPI_ISL_523542\| | ···················· | ·A·················· | ···················· | ···················· | ···················· | ··A················· |
| G | 7810 hCoV-19/Netherlands/UT-EMC-66/2020\|EPI_ISL_523543\| | ···················· | ···················· | ···················· | ···················· | ···················· | ···················· |
| G | 7811 hCoV-19/Netherlands/UT-EMC-67/2020\|EPI_ISL_523544\| | ···················· | ·A·················· | ···················· | ···················· | ···················· | ···················· |
| G | 7812 hCoV-19/Netherlands/UT-EMC-68/2020\|EPI_ISL_523545\| | ···················· | ···················· | ···················· | ···················· | ···················· | ···················· |
| G | 7813 hCoV-19/Netherlands/UT-EMC-69/2020\|EPI_ISL_523546\| | ···················· | ·A·················· | ···················· | ···················· | ···················· | ···················· |
| G | 7814 hCoV-19/Netherlands/ZE-EMC-48/2020\|EPI_ISL_523547\| | ···················· | ···················· | ···················· | ···················· | ···················· | ···················· |
| G | 7815 hCoV-19/Netherlands/ZE-EMC-50/2020\|EPI_ISL_523549\| | ···················· | ···················· | ···················· | ···················· | ···················· | ···················· |
| G | 7816 hCoV-19/Netherlands/ZE-EMC-51/2020\|EPI_ISL_523550\| | ···················· | ···················· | ···················· | ···················· | ···················· | ···················· |
| G | 7817 hCoV-19/Netherlands/ZE-EMC-52/2020\|EPI_ISL_523551\| | ···················· | ···················· | ···················· | ···················· | ···················· | ···················· |
| G | 7818 hCoV-19/Netherlands/ZE-EMC-53/2020\|EPI_ISL_523552\| | ···················· | ···················· | ···················· | ···················· | ···················· | ···················· |
| G | 7819 hCoV-19/Netherlands/ZE-EMC-54/2020\|EPI_ISL_523553\| | ···················· | ···················· | ···················· | ···················· | ···················· | ···················· |
| G | 7820 hCoV-19/Netherlands/ZE-EMC-55/2020\|EPI_ISL_523554\| | ···················· | ···················· | ···················· | ···················· | ···················· | ···················· |
| G | 7821 hCoV-19/Netherlands/ZE-EMC-56/2020\|EPI_ISL_523555\| | ···················· | ···················· | ···················· | ···················· | ···················· | ···················· |
| G | 7822 hCoV-19/Netherlands/ZE-EMC-57/2020\|EPI_ISL_523556\| | ···················· | ···················· | ···················· | ···················· | ···················· | ···················· |
| G | 7823 hCoV-19/Netherlands/ZE-EMC-58/2020\|EPI_ISL_523557\| | ···················· | ···················· | ···················· | ···················· | ···················· | ···················· |
| G | 7824 hCoV-19/Netherlands/ZH-EMC-228/2020\|EPI_ISL_523570 | ···················· | ···················· | ···················· | ···················· | ···················· | ···················· |
| G | 7825 hCoV-19/Netherlands/ZH-EMC-229/2020\|EPI_ISL_523571 | ···················· | ···················· | ···················· | ···················· | ···················· | ···················· |
| G | 7826 hCoV-19/Netherlands/ZH-EMC-233/2020\|EPI_ISL_523575 | ···················· | ···················· | ···················· | ···················· | ···················· | ···················· |
| G | 7827 hCoV-19/Netherlands/ZH-EMC-238/2020\|EPI_ISL_523580 | ···················· | ···················· | ···················· | ···················· | ···················· | ···················· |
| G | 7828 hCoV-19/Netherlands/ZH-EMC-241/2020\|EPI_ISL_523583 | ···················· | ···················· | ···················· | ···················· | ···················· | ···················· |
| G | 7829 hCoV-19/Netherlands/ZH-EMC-242/2020\|EPI_ISL_523584 | ···················· | ···················· | ···················· | ···················· | ···················· | ···················· |
| G | 7830 hCoV-19/Netherlands/ZH-EMC-243/2020\|EPI_ISL_523585 | ···················· | ···················· | ···················· | ···················· | ···················· | ···················· |
| G | 7831 hCoV-19/Netherlands/ZH-EMC-244/2020\|EPI_ISL_523586 | ···················· | ···················· | ···················· | ···················· | ···················· | ···················· |
| G | 7832 hCoV-19/Netherlands/ZH-EMC-245/2020\|EPI_ISL_523587 | ···················· | ···················· | ···················· | ···················· | ···················· | ···················· |
| G | 7833 hCoV-19/Netherlands/ZH-EMC-246/2020\|EPI_ISL_523588 | ···················· | ···················· | ···················· | ···················· | ···················· | ···················· |
| G | 7834 hCoV-19/Netherlands/ZH-EMC-249/2020\|EPI_ISL_523591 | ···················· | ···················· | ···················· | ···················· | ···················· | ···················· |
| G | 7835 hCoV-19/Netherlands/ZH-EMC-250/2020\|EPI_ISL_523592 | ···················· | ···················· | ···················· | ···················· | ···················· | ···················· |
| G | 7836 hCoV-19/Netherlands/ZH-EMC-251/2020\|EPI_ISL_523593 | ···················· | ···················· | ···················· | ···················· | ···················· | ···················· |
| G | 7837 hCoV-19/Netherlands/ZH-EMC-254/2020\|EPI_ISL_523596 | ···················· | ···················· | ···················· | ···················· | ···················· | ···················· |
| G | 7838 hCoV-19/Netherlands/ZH-EMC-255/2020\|EPI_ISL_523597 | ···················· | ···················· | ···················· | ···················· | ···················· | ···················· |
| G | 7839 hCoV-19/Netherlands/ZH-EMC-256/2020\|EPI_ISL_523598 | ···················· | ···················· | ···················· | ···················· | ···················· | ···················· |
| G | 7840 hCoV-19/Netherlands/ZH-EMC-257/2020\|EPI_ISL_523599 | ···················· | ···················· | ···················· | ···················· | ···················· | ···················· |
| G | 7841 hCoV-19/Netherlands/ZH-EMC-360/2020\|EPI_ISL_523700 | ···················· | ···················· | ···················· | ···················· | ···················· | ···················· |
| G | 7842 hCoV-19/Netherlands/ZH-EMC-361/2020\|EPI_ISL_523701 | ···················· | ···················· | ···················· | ···················· | ···················· | ···················· |
| G | 7843 hCoV-19/Netherlands/ZH-EMC-362/2020\|EPI_ISL_523702 | ···················· | ···················· | ···················· | ···················· | ···················· | ···················· |
| G | 7844 hCoV-19/Netherlands/ZH-EMC-363/2020\|EPI_ISL_523703 | ···················· | ···················· | ···················· | ···················· | ···················· | ···················· |
| G | 7845 hCoV-19/Netherlands/ZH-EMC-364/2020\|EPI_ISL_523704 | ···················· | ···················· | ···················· | ···················· | ···················· | ···················· |
| G | 7846 hCoV-19/Netherlands/ZH-EMC-365/2020\|EPI_ISL_523705 | ···················· | ···················· | ···················· | ···················· | ···················· | ···················· |
| G | 7847 hCoV-19/Netherlands/ZH-EMC-366/2020\|EPI_ISL_523706 | ···················· | ···················· | ···················· | ···················· | ···················· | ···················· |
| G | 7848 hCoV-19/Netherlands/ZH-EMC-367/2020\|EPI_ISL_523707 | ···················· | ···················· | ···················· | ···················· | ···················· | ···················· |
| G | 7849 hCoV-19/Netherlands/ZH-EMC-368/2020\|EPI_ISL_523708 | ···················· | ···················· | ···················· | ···················· | ···················· | ···················· |
| G | 7850 hCoV-19/Netherlands/ZH-EMC-369/2020\|EPI_ISL_523709 | ···················· | ···················· | ···················· | ···················· | ···················· | ···················· |
| G | 7851 hCoV-19/Netherlands/ZH-EMC-370/2020\|EPI_ISL_523710 | ···················· | ···················· | ···················· | ···················· | ···················· | ···················· |
| G | 7852 hCoV-19/Netherlands/ZH-EMC-371/2020\|EPI_ISL_523711 | ···················· | ···················· | ···················· | ···················· | ···················· | ···················· |
| G | 7853 hCoV-19/Brazil/PB-IEC-161853/2020\|EPI_ISL_524801\|2 | ···················· | ···················· | ···················· | ···················· | ···················· | ···················· |
| G | 7854 hCoV-19/USA/UT-UPHL-2008769/2020\|EPI_ISL_524802\|20 | ···················· | ···················· | ···················· | ···················· | ···················· | ···················· |
| G | 7855 hCoV-19/USA/UT-UPHL-2008771/2020\|EPI_ISL_524804\|20 | ···················· | ···················· | ···················· | ···················· | ···················· | ···················· |
| G | 7856 hCoV-19/USA/UT-UPHL-2008772/2020\|EPI_ISL_524805\|20 | ···················· | ···················· | ···················· | ···················· | ···················· | ···················· |
| G | 7857 hCoV-19/USA/UT-UPHL-2008774/2020\|EPI_ISL_524806\|20 | ···················· | ·A·················· | ···················· | ···················· | ···················· | ···················· |
| G | 7858 hCoV-19/USA/UT-UPHL-2008775/2020\|EPI_ISL_524807\|20 | ···················· | ···················· | ···················· | ···················· | ···················· | ···················· |
| G | 7859 hCoV-19/USA/UT-UPHL-2008777/2020\|EPI_ISL_524808\|20 | ···················· | ···················· | ···················· | ···················· | ···················· | ···················· |
| G | 7860 hCoV-19/USA/UT-UPHL-2008778/2020\|EPI_ISL_524809\|20 | ···················· | ···················· | ···················· | ···················· | ···················· | ···················· |
| G | 7861 hCoV-19/USA/UT-UPHL-2008779/2020\|EPI_ISL_524810\|20 | ···················· | ···················· | ···················· | ···················· | ···················· | ···················· |
| G | 7862 hCoV-19/USA/UT-UPHL-2008783/2020\|EPI_ISL_524811\|20 | ···················· | ·A·················· | ···················· | ···················· | ···················· | ···················· |
| G | 7863 hCoV-19/USA/UT-UPHL-2008785/2020\|EPI_ISL_524812\|20 | ···················· | ···················· | ···················· | ···················· | ···················· | ···················· |
| G | 7864 hCoV-19/USA/UT-UPHL-2008786/2020\|EPI_ISL_524813\|20 | ···················· | ···················· | ···················· | ···················· | ···················· | ···················· |
| G | 7865 hCoV-19/USA/UT-UPHL-2008787/2020\|EPI_ISL_524814\|20 | ···················· | ···················· | ···················· | ···················· | ···················· | ···················· |
| G | 7866 hCoV-19/USA/UT-UPHL-2008788/2020\|EPI_ISL_524815\|20 | ···················· | ···················· | ···················· | ···················· | ········A··········· | ···················· |
| G | 7867 hCoV-19/USA/UT-UPHL-2008790/2020\|EPI_ISL_524817\|20 | ···················· | ···················· | ···················· | ···················· | ···················· | ···················· |
| G | 7868 hCoV-19/USA/UT-UPHL-2008791/2020\|EPI_ISL_524818\|20 | ···················· | ···················· | ···················· | ···················· | ···················· | ···················· |
| G | 7869 hCoV-19/USA/UT-UPHL-2008792/2020\|EPI_ISL_524819\|20 | ···················· | ···················· | ···················· | ···················· | ···················· | ···················· |
| G | 7870 hCoV-19/USA/UT-UPHL-2008794/2020\|EPI_ISL_524821\|20 | ···················· | ···················· | ···················· | ···················· | ···················· | ···················· |

| | | | | | | | | |
|---|---|---|---|---|---|---|---|---|
| G | 7871 | hCoV-19/USA/UT-UPHL-2008795/2020\|EPI_ISL_524822\|20 | ···················· | ···················· | ···················· | ···················· | ···················· | ···················· |
| G | 7872 | hCoV-19/USA/UT-UPHL-2008798/2020\|EPI_ISL_524824\|20 | ···················· | ···················· | ···················· | ···················· | ···················· | ···················· |
| G | 7873 | hCoV-19/USA/UT-UPHL-2008799/2020\|EPI_ISL_524825\|20 | ···················· | ···················· | ···················· | ···················· | ···················· | ···················· |
| G | 7874 | hCoV-19/USA/UT-UPHL-2008800/2020\|EPI_ISL_524826\|20 | ···················· | ···················· | ···················· | ···················· | ···················· | ···················· |
| G | 7875 | hCoV-19/USA/UT-UPHL-2008802/2020\|EPI_ISL_524828\|20 | ···················· | ···················· | ···················· | ···················· | ···················· | ···················· |
| G | 7876 | hCoV-19/USA/UT-UPHL-2008803/2020\|EPI_ISL_524829\|20 | ···················· | ···················· | ···················· | ···················· | ···················· | ···················· |
| G | 7877 | hCoV-19/USA/UT-UPHL-2008804/2020\|EPI_ISL_524830\|20 | ···················· | ·A·················· | ···················· | ···················· | ···················· | ···················· |
| G | 7878 | hCoV-19/USA/UT-UPHL-2008805/2020\|EPI_ISL_524831\|20 | ···················· | ···················· | ···················· | ···················· | ···················· | ···················· |
| G | 7879 | hCoV-19/USA/UT-UPHL-2008806/2020\|EPI_ISL_524832\|20 | ···················· | ···················· | ···················· | ···················· | ···················· | ···················· |
| G | 7880 | hCoV-19/USA/UT-UPHL-2008807/2020\|EPI_ISL_524833\|20 | ···················· | ···················· | ···················· | ···················· | ···················· | ···················· |
| G | 7881 | hCoV-19/USA/UT-UPHL-2008808/2020\|EPI_ISL_524834\|20 | ···················· | ···················· | ···················· | ···················· | ···················· | ···················· |
| G | 7882 | hCoV-19/USA/UT-UPHL-2008809/2020\|EPI_ISL_524835\|20 | ···················· | ···················· | ···················· | ···················· | ···················· | ···················· |
| G | 7883 | hCoV-19/USA/UT-UPHL-2008810/2020\|EPI_ISL_524836\|20 | ···················· | ···················· | ···················· | ···················· | ···················· | ···················· |
| G | 7884 | hCoV-19/USA/UT-UPHL-2008812/2020\|EPI_ISL_524837\|20 | ···················· | ···················· | ···················· | ···················· | ···················· | ···················· |
| G | 7885 | hCoV-19/USA/UT-UPHL-2008813/2020\|EPI_ISL_524838\|20 | ···················· | ···················· | ···················· | ···················· | ···················· | ···················· |
| G | 7886 | hCoV-19/USA/UT-UPHL-2008816/2020\|EPI_ISL_524840\|20 | ···················· | ···················· | ···················· | ···················· | ·········A·········· | ···················· |
| G | 7887 | hCoV-19/USA/UT-UPHL-2008818/2020\|EPI_ISL_524841\|20 | ···················· | ···················· | ···················· | ···················· | ···················· | ···················· |
| G | 7888 | hCoV-19/USA/UT-UPHL-2008821/2020\|EPI_ISL_524842\|20 | ···················· | ···················· | ···················· | ···················· | ···················· | ···················· |
| G | 7889 | hCoV-19/USA/UT-UPHL-2008822/2020\|EPI_ISL_524843\|20 | ···················· | ···················· | ···················· | ···················· | ···················· | ···················· |
| G | 7890 | hCoV-19/USA/UT-UPHL-2008823/2020\|EPI_ISL_524844\|20 | ···················· | ···················· | ···················· | ···················· | ···················· | ···················· |
| G | 7891 | hCoV-19/USA/UT-UPHL-2008824/2020\|EPI_ISL_524845\|20 | ···················· | ···················· | ···················· | ···················· | ···················· | ···················· |
| G | 7892 | hCoV-19/USA/UT-UPHL-2008825/2020\|EPI_ISL_524846\|20 | ···················· | ···················· | ···················· | ···················· | ···················· | ···················· |
| G | 7893 | hCoV-19/USA/UT-UPHL-2008828/2020\|EPI_ISL_524848\|20 | ···················· | ···················· | ···················· | ···················· | ···················· | ···················· |
| G | 7894 | hCoV-19/USA/UT-UPHL-2008829/2020\|EPI_ISL_524849\|20 | ···················· | ···················· | ···················· | ···················· | ···················· | ···················· |
| G | 7895 | hCoV-19/USA/UT-UPHL-2008830/2020\|EPI_ISL_524850\|20 | ···················· | ···················· | ···················· | ···················· | ···················· | ···················· |
| G | 7896 | hCoV-19/USA/UT-UPHL-2008831/2020\|EPI_ISL_524851\|20 | ···················· | ···················· | ···················· | ···················· | ···················· | ···················· |
| G | 7897 | hCoV-19/USA/UT-UPHL-2008832/2020\|EPI_ISL_524852\|20 | ···················· | ···················· | A··················· | ···················· | ···················· | ···················· |
| G | 7898 | hCoV-19/USA/UT-UPHL-2008833/2020\|EPI_ISL_524853\|20 | ···················· | ···················· | ···················· | ···················· | ·········A·········· | ···················· |
| G | 7899 | hCoV-19/USA/UT-UPHL-2008836/2020\|EPI_ISL_524855\|20 | ···················· | ···················· | ···················· | ···················· | ·········A·········· | ···················· |
| G | 7900 | hCoV-19/USA/UT-UPHL-2008837/2020\|EPI_ISL_524856\|20 | ···················· | ···················· | ···················· | ···················· | ···················· | ···················· |
| G | 7901 | hCoV-19/USA/UT-UPHL-2008839/2020\|EPI_ISL_524857\|20 | ···················· | ···················· | ···················· | ···················· | ···················· | ···················· |
| G | 7902 | hCoV-19/USA/UT-UPHL-2008841/2020\|EPI_ISL_524858\|20 | ···················· | ···················· | ···················· | ···················· | ···················· | ···················· |
| G | 7903 | hCoV-19/USA/UT-UPHL-2008842/2020\|EPI_ISL_524859\|20 | ···················· | ·A·················· | ···················· | ···················· | ···················· | ···················· |
| G | 7904 | hCoV-19/USA/UT-UPHL-2008844/2020\|EPI_ISL_524861\|20 | ···················· | ···················· | ···················· | ···················· | ···················· | ···················· |
| G | 7905 | hCoV-19/USA/UT-UPHL-2008845/2020\|EPI_ISL_524862\|20 | ···················· | ···················· | ···················· | ···················· | ···················· | ···················· |
| G | 7906 | hCoV-19/USA/UT-UPHL-2008846/2020\|EPI_ISL_524863\|20 | ···················· | ···················· | ···················· | ···················· | ···················· | ···················· |
| G | 7907 | hCoV-19/USA/UT-UPHL-2008847/2020\|EPI_ISL_524864\|20 | ···················· | ···················· | ···················· | ···················· | ···················· | ···················· |
| G | 7908 | hCoV-19/USA/UT-UPHL-2008851/2020\|EPI_ISL_524867\|20 | ···················· | ···················· | ···················· | ···················· | ···················· | ···················· |
| G | 7909 | hCoV-19/USA/UT-UPHL-2008852/2020\|EPI_ISL_524868\|20 | ···················· | ···················· | ···················· | ···················· | ···················· | ···················· |
| G | 7910 | hCoV-19/USA/UT-UPHL-2008853/2020\|EPI_ISL_524869\|20 | ···················· | ···················· | ···················· | ···················· | ···················· | ···················· |
| G | 7911 | hCoV-19/USA/UT-UPHL-2008854/2020\|EPI_ISL_524870\|20 | ···················· | ···················· | ···················· | ···················· | ···················· | ···················· |
| G | 7912 | hCoV-19/USA/UT-UPHL-2008855/2020\|EPI_ISL_524871\|20 | ···················· | ···················· | ···················· | ···················· | ···················· | ···················· |
| G | 7913 | hCoV-19/USA/UT-UPHL-2008856/2020\|EPI_ISL_524872\|20 | ···················· | ···················· | ···················· | ···················· | ···················· | ···················· |
| G | 7914 | hCoV-19/USA/MD-MDH-0145/2020\|EPI_ISL_524873\|2020-0 | ···················· | ···················· | ···················· | ···················· | ···················· | ···················· |
| G | 7915 | hCoV-19/USA/MD-MDH-0147/2020\|EPI_ISL_524875\|2020-0 | ···················· | ···················· | ···················· | ···················· | ···················· | ···················· |
| G | 7916 | hCoV-19/USA/MD-MDH-0148/2020\|EPI_ISL_524876\|2020-0 | ···················· | ···················· | ···················· | ···················· | ···················· | ···················· |
| G | 7917 | hCoV-19/USA/MD-MDH-0149/2020\|EPI_ISL_524877\|2020-0 | ···················· | ···················· | ···················· | ···················· | ···················· | ···················· |
| G | 7918 | hCoV-19/USA/MD-MDH-0150/2020\|EPI_ISL_524878\|2020-0 | ···················· | ···················· | ···················· | ···················· | ···················· | ···················· |
| G | 7919 | hCoV-19/USA/MD-MDH-0151/2020\|EPI_ISL_524879\|2020-0 | ···················· | ···················· | ···················· | ···················· | ···················· | ···················· |
| G | 7920 | hCoV-19/USA/MD-MDH-0152/2020\|EPI_ISL_524880\|2020-0 | ···················· | ···················· | ···················· | ···················· | ···················· | ···················· |
| G | 7921 | hCoV-19/USA/MD-MDH-0153/2020\|EPI_ISL_524881\|2020-0 | ···················· | ···················· | ···················· | ···················· | ···················· | ···················· |
| G | 7922 | hCoV-19/USA/MD-MDH-0154/2020\|EPI_ISL_524882\|2020-0 | ···················· | ···················· | ···················· | ···················· | ···················· | ···················· |
| G | 7923 | hCoV-19/USA/MD-MDH-0157/2020\|EPI_ISL_524884\|2020-0 | ···················· | ···················· | ···················· | ···················· | ···················· | ···················· |
| G | 7924 | hCoV-19/USA/MD-MDH-0158/2020\|EPI_ISL_524885\|2020-0 | ···················· | ···················· | ···················· | ···················· | ···················· | ···················· |
| G | 7925 | hCoV-19/USA/MD-MDH-0159/2020\|EPI_ISL_524886\|2020-0 | ···················· | ······A············· | ···················· | ···················· | ···················· | ···················· |
| G | 7926 | hCoV-19/USA/MD-MDH-0160/2020\|EPI_ISL_524887\|2020-0 | ···················· | ···················· | ···················· | ···················· | ···················· | ···················· |
| G | 7927 | hCoV-19/USA/MD-MDH-0161/2020\|EPI_ISL_524888\|2020-0 | ···················· | ···················· | ···················· | ···················· | ···················· | ···················· |
| G | 7928 | hCoV-19/USA/AK-PHL1895/2020\|EPI_ISL_528513\|2020-07 | ···················· | ···················· | ···················· | ···················· | ···················· | ···················· |
| G | 7929 | hCoV-19/USA/AK-PHL803/2020\|EPI_ISL_528526\|2020-07- | ···················· | ···················· | ···················· | ···················· | ···················· | ···················· |
| G | 7930 | hCoV-19/India/TG-CDFD-C2169/2020\|EPI_ISL_528576\|20 | ···················· | ···················· | ···················· | ···················· | ···················· | ······N············· |
| G | 7931 | hCoV-19/India/TG-CDFD-C2529/2020\|EPI_ISL_528581\|20 | ···················· | ···················· | ···················· | ···················· | ···················· | ···················· |
| G | 7932 | hCoV-19/India/TG-CDFD-C2910/2020\|EPI_ISL_528586\|20 | ···················· | ···················· | ···················· | ···················· | ···················· | ···················· |
| G | 7934 | hCoV-19/USA/MI-MDHHS-SC21711/2020\|EPI_ISL_529830\|2 | ···················· | ···················· | ···················· | ···················· | ···················· | ···················· |
| G | 7937 | hCoV-19/Peru/CAJ-INS-061/2020\|EPI_ISL_517770\|2020- | ···················· | ···················· | ···················· | ···················· | ···················· | ···················· |
| G | 7938 | hCoV-19/USA/FL-BPHL-0881/2020\|EPI_ISL_517772\|2020- | ···················· | ···················· | ···················· | ···················· | ···················· | ···················· |
| G | 7939 | hCoV-19/USA/FL-BPHL-0888/2020\|EPI_ISL_517778\|2020- | ···················· | ···················· | ···················· | ···················· | ···················· | ···················· |
| G | 7940 | hCoV-19/USA/FL-BPHL-0900/2020\|EPI_ISL_517790\|2020- | ···················· | ···················· | ···················· | ···················· | ···················· | ···················· |
| G | 7941 | hCoV-19/USA/FL-BPHL-0901/2020\|EPI_ISL_517791\|2020- | ···················· | ···················· | ···················· | ···················· | ···················· | ···················· |
| G | 7942 | hCoV-19/USA/FL-BPHL-0902/2020\|EPI_ISL_517792\|2020- | ···················· | ···················· | ···················· | ···················· | ···················· | ···················· |
| G | 7943 | hCoV-19/USA/FL-BPHL-0904/2020\|EPI_ISL_517794\|2020- | ···················· | ···················· | ···················· | ···················· | ···················· | ···················· |
| G | 7944 | hCoV-19/USA/FL-BPHL-0905/2020\|EPI_ISL_517795\|2020- | ···················· | ········A··········· | ···················· | ···················· | ···················· | ···················· |
| G | 7947 | hCoV-19/Australia/VIC2382/2020\|EPI_ISL_522101\|2020 | ···················· | ···················· | ···················· | ···················· | ···················· | ···················· |
| G | 7948 | hCoV-19/Australia/VIC2384/2020\|EPI_ISL_522102\|2020 | ···················· | ···················· | ···················· | ···················· | ···················· | ···················· |
| G | 7949 | hCoV-19/Australia/VIC2385/2020\|EPI_ISL_522103\|2020 | ···················· | ···················· | ···················· | ···················· | ···················· | ···················· |
| G | 7950 | hCoV-19/Australia/VIC2386/2020\|EPI_ISL_522104\|2020 | ···················· | ···················· | ···················· | ···················· | ···················· | ···················· |
| G | 7951 | hCoV-19/Australia/VIC2534/2020\|EPI_ISL_522227\|2020 | ···················· | ···················· | ···················· | ···················· | ···················· | ···················· |
| G | 7952 | hCoV-19/Australia/VIC2535/2020\|EPI_ISL_522228\|2020 | ···················· | ···················· | ···················· | ···················· | ···················· | ···················· |
| G | 7953 | hCoV-19/Australia/VIC2536/2020\|EPI_ISL_522229\|2020 | ···················· | ···················· | ···················· | ···················· | ···················· | ···················· |
| G | 7954 | hCoV-19/Australia/VIC2537/2020\|EPI_ISL_522230\|2020 | ···················· | ···················· | ···················· | ···················· | ···················· | ···················· |
| G | 7955 | hCoV-19/Australia/VIC2538/2020\|EPI_ISL_522231\|2020 | ···················· | ···················· | ···················· | ···················· | ···················· | ···················· |

| | | | | | | | |
|---|---|---|---|---|---|---|---|
| G | 7956 hCoV-19/Australia/VIC2546/2020\|EPI_ISL_522233\|2020 | ···················· | ···················· | ···················· | ···················· | ···················· | ···················· |
| G | 7957 hCoV-19/Australia/VIC2547/2020\|EPI_ISL_522234\|2020 | ···················· | ···················· | ···················· | ···················· | ···················· | ···················· |
| G | 7958 hCoV-19/Australia/VIC2548/2020\|EPI_ISL_522235\|2020 | ···················· | ···················· | ···················· | ···················· | N··················· | ···················· |
| G | 7959 hCoV-19/Australia/VIC2549/2020\|EPI_ISL_522236\|2020 | ···················· | ···················· | ···················· | ···················· | ···················· | ···················· |
| G | 7960 hCoV-19/Australia/VIC2550/2020\|EPI_ISL_522237\|2020 | ···················· | ···················· | ···················· | ···················· | ···················· | ···················· |
| G | 7961 hCoV-19/Australia/VIC2551/2020\|EPI_ISL_522238\|2020 | ···················· | ···················· | ···················· | ···················· | ···················· | ···················· |
| G | 7962 hCoV-19/Australia/VIC2553/2020\|EPI_ISL_522239\|2020 | ···················· | ···················· | ···················· | ···················· | ···················· | ···················· |
| G | 7963 hCoV-19/Australia/VIC2557/2020\|EPI_ISL_522241\|2020 | ···················· | ···················· | ···················· | ···················· | ···················· | ···················· |
| G | 7964 hCoV-19/Australia/VIC2558/2020\|EPI_ISL_522242\|2020 | ···················· | ···················· | ···················· | ···················· | ···················· | ···················· |
| G | 7965 hCoV-19/Australia/VIC2561/2020\|EPI_ISL_522245\|2020 | ···················· | ···················· | ···················· | ···················· | ···················· | ···················· |
| G | 7966 hCoV-19/Australia/VIC2569/2020\|EPI_ISL_522247\|2020 | ···················· | ···················· | ···················· | ···················· | ···················· | ···················· |
| G | 7967 hCoV-19/Netherlands/NH-EMC-41/2020\|EPI_ISL_523410\| | ···················· | ···················· | ···················· | ···················· | ···················· | ···················· |
| G | 7968 hCoV-19/Netherlands/NH-EMC-46/2020\|EPI_ISL_523415\| | ···················· | ···················· | ···················· | ···················· | ···················· | ···················· |
| G | 7969 hCoV-19/Netherlands/NH-EMC-47/2020\|EPI_ISL_523416\| | ···················· | ···················· | ···················· | ···················· | ···················· | ···················· |
| G | 7970 hCoV-19/Netherlands/NH-EMC-48/2020\|EPI_ISL_523417\| | ···················· | ···················· | ···················· | ···················· | ···················· | ···················· |
| G | 7971 hCoV-19/Netherlands/OV-EMC-36/2020\|EPI_ISL_523420\| | ···················· | ···················· | ···················· | ···················· | ···················· | ···················· |
| G | 7972 hCoV-19/Netherlands/OV-EMC-37/2020\|EPI_ISL_523421\| | ···················· | ···················· | ···················· | ···················· | ···················· | ···················· |
| G | 7973 hCoV-19/Netherlands/OV-EMC-38/2020\|EPI_ISL_523422\| | ···················· | ···················· | ···················· | ···················· | ···················· | ···················· |
| G | 7974 hCoV-19/Netherlands/OV-EMC-39/2020\|EPI_ISL_523423\| | ···················· | ···················· | ···················· | ···················· | ···················· | ···················· |
| G | 7975 hCoV-19/Netherlands/OV-EMC-40/2020\|EPI_ISL_523424\| | ···················· | ···················· | ···················· | ···················· | ···················· | ···················· |
| G | 7976 hCoV-19/Netherlands/OV-EMC-41/2020\|EPI_ISL_523425\| | ···················· | ···················· | ···················· | ···················· | ···················· | ···················· |
| G | 7977 hCoV-19/Netherlands/OV-EMC-42/2020\|EPI_ISL_523426\| | ···················· | ···················· | ···················· | ···················· | ···················· | ···················· |
| G | 7978 hCoV-19/Netherlands/OV-EMC-43/2020\|EPI_ISL_523427\| | ···················· | ···················· | ···················· | ···················· | ···················· | ···················· |
| G | 7979 hCoV-19/Netherlands/OV-EMC-44/2020\|EPI_ISL_523428\| | ···················· | ···················· | ···················· | ···················· | ···················· | ···················· |
| G | 7980 hCoV-19/Netherlands/OV-EMC-45/2020\|EPI_ISL_523429\| | ···················· | ···················· | ···················· | ···················· | ···················· | ···················· |
| G | 7981 hCoV-19/Netherlands/OV-EMC-46/2020\|EPI_ISL_523430\| | ···················· | ···················· | ···················· | ···················· | ···················· | ···················· |
| G | 7982 hCoV-19/Netherlands/OV-EMC-47/2020\|EPI_ISL_523431\| | ···················· | ···················· | ···················· | ···················· | ···················· | ···················· |
| G | 7983 hCoV-19/Netherlands/OV-EMC-48/2020\|EPI_ISL_523432\| | ···················· | ···················· | ···················· | ···················· | ···················· | ···················· |
| G | 7984 hCoV-19/Netherlands/OV-EMC-49/2020\|EPI_ISL_523433\| | ···················· | ···················· | ···················· | ···················· | ···················· | ···················· |
| G | 7985 hCoV-19/Netherlands/OV-EMC-50/2020\|EPI_ISL_523434\| | ···················· | ···················· | ···················· | ···················· | ···················· | ···················· |
| G | 7986 hCoV-19/Netherlands/OV-EMC-51/2020\|EPI_ISL_523435\| | ···················· | ···········A········ | ···················· | ···················· | ···················· | ···················· |
| G | 7987 hCoV-19/Netherlands/OV-EMC-52/2020\|EPI_ISL_523436\| | ···················· | ···········A········ | ···················· | ···················· | ···················· | ···················· |
| G | 7988 hCoV-19/Netherlands/ZE-EMC-59/2020\|EPI_ISL_523558\| | ···················· | ···················· | ···················· | ···················· | ···················· | ···················· |
| G | 7989 hCoV-19/Netherlands/ZE-EMC-60/2020\|EPI_ISL_523559\| | ···················· | ···················· | ···················· | ···················· | ···················· | ···················· |
| G | 7990 hCoV-19/Netherlands/ZE-EMC-61/2020\|EPI_ISL_523560\| | ···················· | ···················· | ···················· | ···················· | ···················· | ···················· |
| G | 7991 hCoV-19/Netherlands/ZE-EMC-64/2020\|EPI_ISL_523563\| | ···················· | ···················· | ···················· | ···················· | ···················· | ···················· |
| G | 7992 hCoV-19/Netherlands/ZE-EMC-65/2020\|EPI_ISL_523564\| | ···················· | ···················· | ···················· | ···················· | ···················· | ···················· |
| G | 7993 hCoV-19/Netherlands/ZE-EMC-66/2020\|EPI_ISL_523565\| | ···················· | ···················· | ···················· | ···················· | ···················· | ···················· |
| G | 7994 hCoV-19/Netherlands/ZE-EMC-67/2020\|EPI_ISL_523566\| | ···················· | ···················· | ···················· | ···················· | ···················· | ···················· |
| G | 7995 hCoV-19/Netherlands/ZE-EMC-69/2020\|EPI_ISL_523568\| | ···················· | ···················· | ···················· | ···················· | ···················· | ···················· |
| G | 7996 hCoV-19/Netherlands/ZH-EMC-227/2020\|EPI_ISL_523569 | ···················· | ···················· | ···················· | ···················· | ···················· | ···················· |
| G | 7997 hCoV-19/Netherlands/ZH-EMC-230/2020\|EPI_ISL_523572 | ···················· | ···················· | ···················· | ···················· | ···················· | ···················· |
| G | 7998 hCoV-19/Netherlands/ZH-EMC-231/2020\|EPI_ISL_523573 | ···················· | ···················· | ···················· | ···················· | ···················· | ···················· |
| G | 7999 hCoV-19/Netherlands/ZH-EMC-234/2020\|EPI_ISL_523576 | ···················· | ···················· | ···················· | ···················· | ···················· | ···················· |
| G | 8000 hCoV-19/Netherlands/ZH-EMC-235/2020\|EPI_ISL_523577 | ···················· | ···················· | ···················· | ···················· | ···················· | ···················· |
| G | 8001 hCoV-19/Netherlands/ZH-EMC-236/2020\|EPI_ISL_523578 | ···················· | ···················· | ···················· | ···················· | ···················· | ···················· |
| G | 8002 hCoV-19/Netherlands/ZH-EMC-237/2020\|EPI_ISL_523579 | ···················· | ···················· | ···················· | ···················· | ···················· | ···················· |
| G | 8003 hCoV-19/India/GJ-GBRC-383a/2020\|EPI_ISL_524742\|202 | ···················· | ·A·················· | ···················· | ···················· | ···················· | ··A················· |
| G | 8004 hCoV-19/India/GJ-GBRC-385/2020\|EPI_ISL_524746\|2020 | ···················· | ···················· | ···················· | ···················· | ···················· | ···················· |
| G | 8005 hCoV-19/India/GJ-GBRC-386a/2020\|EPI_ISL_524747\|202 | ···················· | ·A·················· | ···················· | ···················· | ···················· | ··A················· |
| G | 8006 hCoV-19/India/GJ-GBRC-386b/2020\|EPI_ISL_524748\|202 | ···················· | ·A·················· | ···················· | ···················· | ···················· | ··A················· |
| G | 8007 hCoV-19/India/GJ-GBRC-387a/2020\|EPI_ISL_524749\|202 | ···················· | ···················· | ···················· | ···················· | ···················· | ···················· |
| G | 8008 hCoV-19/India/GJ-GBRC-387b/2020\|EPI_ISL_524750\|202 | ···················· | ···················· | ···················· | ···················· | ···················· | ···················· |
| G | 8009 hCoV-19/India/GJ-GBRC-388/2020\|EPI_ISL_524751\|2020 | ···················· | ·A·················· | ···················· | ···················· | ···················· | ··A················· |
| G | 8010 hCoV-19/India/GJ-GBRC-389a/2020\|EPI_ISL_524752\|202 | ···················· | ···················· | ···················· | ···················· | ···················· | ···················· |
| G | 8011 hCoV-19/India/GJ-GBRC-389b/2020\|EPI_ISL_524753\|202 | ········T··········· | ···················· | ···················· | ···················· | ···················· | ···················· |
| G | 8012 hCoV-19/India/GJ-GBRC-390a/2020\|EPI_ISL_524754\|202 | ···················· | ·A·················· | ···················· | ···················· | ···················· | ··A················· |
| G | 8013 hCoV-19/India/GJ-GBRC-390b/2020\|EPI_ISL_524755\|202 | ···················· | ·A·················· | ···················· | ···················· | ···················· | ···················· |
| G | 8014 hCoV-19/India/GJ-GBRC-391a/2020\|EPI_ISL_524756\|202 | ···················· | ·A·················· | ···················· | ···················· | ···················· | ··A················· |
| G | 8015 hCoV-19/India/GJ-GBRC-391b/2020\|EPI_ISL_524757\|202 | ···················· | ·A·················· | ···················· | ···················· | ···················· | ··A················· |
| G | 8016 hCoV-19/India/GJ-GBRC-392a/2020\|EPI_ISL_524758\|202 | ···················· | ···A················ | ···················· | ···················· | ···················· | ···················· |
| G | 8017 hCoV-19/India/GJ-GBRC-392b/2020\|EPI_ISL_524759\|202 | ···················· | ···A················ | ···················· | ···················· | ···················· | ···················· |
| G | 8018 hCoV-19/India/GJ-GBRC-393a/2020\|EPI_ISL_524760\|202 | ···················· | ···················· | ···················· | ···················· | ···················· | ···················· |
| G | 8019 hCoV-19/India/GJ-GBRC-393b/2020\|EPI_ISL_524761\|202 | ···················· | ···················· | ···················· | ···················· | ···················· | ···················· |
| G | 8020 hCoV-19/India/GJ-GBRC-394a/2020\|EPI_ISL_524762\|202 | ···················· | ···················· | ···················· | ···················· | ···················· | ···················· |
| G | 8021 hCoV-19/India/GJ-GBRC-394b/2020\|EPI_ISL_524763\|202 | ···················· | ···················· | ···················· | ···················· | ···················· | ···················· |
| G | 8022 hCoV-19/India/GJ-GBRC-395a/2020\|EPI_ISL_524764\|202 | ···················· | ···················· | ···················· | ···················· | ···················· | ···················· |
| G | 8023 hCoV-19/India/GJ-GBRC-395b/2020\|EPI_ISL_524765\|202 | ············T······· | ···················· | ···················· | ···················· | ···················· | ···················· |
| G | 8024 hCoV-19/India/GJ-GBRC-396/2020\|EPI_ISL_524766\|2020 | ···················· | ·A·················· | ···················· | ···················· | ···················· | ··A················· |
| G | 8025 hCoV-19/USA/UT-08709/2020\|EPI_ISL_524767\|2020-05-1 | ···················· | ···················· | ···················· | ···················· | ···················· | ···················· |
| G | 8026 hCoV-19/USA/MD-MDH-0162/2020\|EPI_ISL_524889\|2020-0 | ···················· | ···················· | ···················· | ···················· | ···················· | ···················· |
| G | 8027 hCoV-19/USA/MD-MDH-0163/2020\|EPI_ISL_524890\|2020-0 | ···················· | ···················· | ···················· | ···················· | ···················· | ···················· |
| G | 8028 hCoV-19/USA/MD-MDH-0164/2020\|EPI_ISL_524891\|2020-0 | ···················· | ···················· | ···················· | ···················· | ···················· | ···················· |
| G | 8029 hCoV-19/USA/MD-MDH-0127/2020\|EPI_ISL_524892\|2020-0 | ···················· | ···················· | ···················· | ···················· | ···················· | ···················· |
| G | 8030 hCoV-19/USA/MD-MDH-0128/2020\|EPI_ISL_524893\|2020-0 | ···················· | ···················· | ···················· | ···················· | ···················· | ···················· |
| G | 8031 hCoV-19/USA/MD-MDH-0129/2020\|EPI_ISL_524894\|2020-0 | ···················· | ···················· | ···················· | ···················· | ···················· | ···················· |
| G | 8032 hCoV-19/USA/MD-MDH-0130/2020\|EPI_ISL_524895\|2020-0 | ···················· | ···················· | ···················· | ···················· | ···················· | ···················· |
| G | 8033 hCoV-19/USA/MD-MDH-0132/2020\|EPI_ISL_524896\|2020-0 | ···················· | ···················· | ···················· | ···················· | ···················· | ···················· |
| G | 8034 hCoV-19/USA/MD-MDH-0133/2020\|EPI_ISL_524897\|2020-0 | ···················· | ···················· | ···················· | ···················· | ···················· | ···················· |
| G | 8035 hCoV-19/USA/MD-MDH-0134/2020\|EPI_ISL_524898\|2020-0 | ···················· | ···················· | ···················· | ···················· | ···················· | ···················· |

| | | | | | | | |
|---|---|---|---|---|---|---|---|
| G | 8036 hCoV-19/USA/MD-MDH-0136/2020\|EPI_ISL_524899\|2020-0 | ···················· | ···················· | ···················· | ···················· | ···················· | ···················· |
| G | 8037 hCoV-19/USA/MD-MDH-0137/2020\|EPI_ISL_524900\|2020-0 | ···················· | ···················· | ···················· | ···················· | ···················· | ···················· |
| G | 8038 hCoV-19/USA/MD-MDH-0138/2020\|EPI_ISL_524901\|2020-0 | ···················· | ···················· | ···················· | ···················· | ···················· | ···················· |
| G | 8039 hCoV-19/USA/UT-UPHL-2009296/2020\|EPI_ISL_524905\|20 | ···················· | ···················· | ···················· | ···················· | ···················· | ···················· |
| G | 8040 hCoV-19/USA/UT-UPHL-2009299/2020\|EPI_ISL_524908\|20 | ···················· | ···················· | ···················· | ···················· | ···················· | ···················· |
| G | 8041 hCoV-19/USA/UT-UPHL-2009304/2020\|EPI_ISL_524910\|20 | ···················· | ···················· | ···················· | ···················· | ···················· | ···················· |
| G | 8042 hCoV-19/USA/UT-UPHL-2009305/2020\|EPI_ISL_524911\|20 | ···················· | ···················· | ···················· | ···················· | ···················· | ···················· |
| G | 8043 hCoV-19/USA/UT-UPHL-2009309/2020\|EPI_ISL_524915\|20 | ···················· | ···················· | ···················· | ···················· | ···················· | ···················· |
| G | 8044 hCoV-19/USA/UT-UPHL-2009311/2020\|EPI_ISL_524917\|20 | ···················· | ···················· | ···················· | ···················· | ···················· | ···················· |
| G | 8045 hCoV-19/USA/UT-UPHL-2009314/2020\|EPI_ISL_524919\|20 | ···················· | ···················· | ···················· | ···················· | ···················· | ···················· |
| G | 8046 hCoV-19/Australia/NSW2716/2020\|EPI_ISL_527179\|2020 | ···················· | ···················· | ···················· | ···················· | ···················· | ···················· |
| G | 8047 hCoV-19/Australia/NSW2717/2020\|EPI_ISL_527180\|2020 | ···················· | ···················· | ···················· | ···················· | ···················· | ···················· |
| G | 8048 hCoV-19/England/20166019504/2020\|EPI_ISL_528400\|20 | ···················· | ···················· | ···················· | ···················· | ···················· | ···················· |
| G | 8049 hCoV-19/England/20166067104/2020\|EPI_ISL_528401\|20 | ···················· | ···················· | ···················· | ···················· | ···················· | ···················· |
| G | 8050 hCoV-19/England/20169038304/2020\|EPI_ISL_528403\|20 | ···················· | ···················· | ···················· | ···················· | ···················· | ···················· |
| G | 8051 hCoV-19/England/20172045304/2020\|EPI_ISL_528404\|20 | ···················· | ···················· | ···················· | ···················· | ···················· | ···················· |
| G | 8052 hCoV-19/England/20172055104/2020\|EPI_ISL_528405\|20 | ···················· | ···················· | ···················· | ···················· | ···················· | ···················· |
| G | 8053 hCoV-19/England/20176001704/2020\|EPI_ISL_528406\|20 | ···················· | ···················· | ···················· | ···················· | ···················· | ···················· |
| G | 8054 hCoV-19/England/20178059804/2020\|EPI_ISL_528407\|20 | ···················· | ···················· | ···················· | ···················· | ···················· | ···················· |
| G | 8055 hCoV-19/England/20186102804/2020\|EPI_ISL_528409\|20 | ···················· | ···················· | ···················· | ···················· | ···················· | ···················· |
| G | 8056 hCoV-19/England/20186106504/2020\|EPI_ISL_528411\|20 | ···················· | ···················· | ···················· | ···················· | ···················· | ···················· |
| G | 8057 hCoV-19/England/20189182904/2020\|EPI_ISL_528413\|20 | ···················· | ···················· | ···················· | ···················· | ···················· | ···················· |
| G | 8058 hCoV-19/England/20190038604/2020\|EPI_ISL_528414\|20 | ···················· | ···················· | ···················· | ···················· | ···················· | ···················· |
| G | 8059 hCoV-19/England/20192003304/2020\|EPI_ISL_528415\|20 | ···················· | ···················· | ···················· | ···················· | ···················· | ···················· |
| G | 8060 hCoV-19/England/20164077504/2020\|EPI_ISL_528430\|20 | ···················· | ···················· | ···················· | ···················· | ···················· | ···················· |
| G | 8061 hCoV-19/England/20179042804/2020\|EPI_ISL_528432\|20 | ···················· | ···················· | ···················· | ···················· | ···················· | ···················· |
| G | 8062 hCoV-19/England/20179044504/2020\|EPI_ISL_528433\|20 | ···················· | ···················· | ···················· | ···················· | ···················· | ···················· |
| G | 8063 hCoV-19/England/20179044904/2020\|EPI_ISL_528434\|20 | ···················· | ···················· | ···················· | ···················· | ···················· | ···················· |
| G | 8064 hCoV-19/England/20179045004/2020\|EPI_ISL_528435\|20 | ···················· | ···················· | ···················· | ···················· | ···················· | ···················· |
| G | 8065 hCoV-19/England/20342000104/2020\|EPI_ISL_528438\|20 | ···················· | ···················· | ···················· | ···················· | ···················· | ···················· |
| G | 8066 hCoV-19/Ireland/D-NVRL-72IRL07177/2020\|EPI_ISL_528 | ···················· | ···················· | ···················· | ···················· | ···················· | ···················· |
| G | 8067 hCoV-19/Ireland/KE-NVRL-72IRL07332/2020\|EPI_ISL_52 | ···················· | ···················· | ···················· | ···················· | ···················· | ···················· |
| G | 8068 hCoV-19/Ireland/KE-NVRL-72IRL07362/2020\|EPI_ISL_52 | ···················· | ···················· | ···················· | ···················· | ···················· | ···················· |
| G | 8069 hCoV-19/Ireland/LK-NVRL-72IRL07419/2020\|EPI_ISL_52 | ···················· | ·A·················· | ···················· | ···················· | ···················· | ··A················· |
| G | 8070 hCoV-19/Ireland/LK-NVRL-72IRL07430/2020\|EPI_ISL_52 | ···················· | ·A·················· | ···················· | ···················· | ···················· | ··A················· |
| G | 8071 hCoV-19/Ireland/LK-NVRL-72IRL07432/2020\|EPI_ISL_52 | ···················· | ·A·················· | ···················· | ···················· | ···················· | ··A················· |
| G | 8072 hCoV-19/Ireland/LK-NVRL-72IRL07454/2020\|EPI_ISL_52 | ···················· | ·A·················· | ···················· | ···················· | ···················· | ··A················· |
| G | 8073 hCoV-19/Ireland/D-NVRL-72IRL07564/2020\|EPI_ISL_528 | ···················· | ·A·················· | ···················· | ···················· | ···················· | ···················· |
| G | 8074 hCoV-19/Ireland/KK-NVRL-72IRL07921/2020\|EPI_ISL_52 | ···················· | ···················· | ···················· | ···················· | ···················· | ···················· |
| G | 8075 hCoV-19/Ireland/KK-NVRL-72IRL07944/2020\|EPI_ISL_52 | ···················· | ···················· | ···················· | ···················· | ···················· | ···················· |
| G | 8076 hCoV-19/Ireland/KK-NVRL-72IRL07961/2020\|EPI_ISL_52 | ···················· | ···················· | ···················· | ···················· | ···················· | ···················· |
| G | 8077 hCoV-19/Ireland/WD-NVRL-72IRL08037/2020\|EPI_ISL_52 | ···················· | ···················· | ···················· | ···················· | ···················· | ···················· |
| G | 8078 hCoV-19/Ireland/MH-NVRL-72IRL08314/2020\|EPI_ISL_52 | ···················· | ···················· | ···················· | ···················· | ···················· | ···················· |
| G | 8079 hCoV-19/Ireland/MH-NVRL-72IRL08381/2020\|EPI_ISL_52 | ···················· | ···················· | ···················· | ···················· | ···················· | ···················· |
| G | 8080 hCoV-19/Ireland/G-NVRL-72IRL10958/2020\|EPI_ISL_528 | ···················· | ···················· | ···················· | ···················· | ···················· | ···················· |
| G | 8081 hCoV-19/Ireland/KK-NVRL-72IRL11130/2020\|EPI_ISL_52 | ···················· | ···················· | ···················· | ···················· | ···················· | ···················· |
| G | 8082 hCoV-19/Ireland/KK-NVRL-72IRL11133/2020\|EPI_ISL_52 | ···················· | ·A·················· | ···················· | ···················· | ···················· | ···················· |
| G | 8083 hCoV-19/Ireland/KK-NVRL-72IRL11143/2020\|EPI_ISL_52 | ···················· | ·A·················· | ···················· | ···················· | ···················· | ···················· |
| G | 8084 hCoV-19/Ireland/KK-NVRL-72IRL11149/2020\|EPI_ISL_52 | ···················· | ···················· | ···················· | ···················· | ···················· | ···················· |
| G | 8085 hCoV-19/Ireland/KK-NVRL-72IRL11375/2020\|EPI_ISL_52 | ···················· | ···················· | ···················· | ···················· | ···················· | ···················· |
| G | 8086 hCoV-19/Ireland/KE-NVRL-72IRL12629/2020\|EPI_ISL_52 | ···················· | ···················· | ···················· | ···················· | ···················· | ···················· |
| G | 8087 hCoV-19/Ireland/KE-NVRL-72IRL12648/2020\|EPI_ISL_52 | ···················· | ···················· | ···················· | ···················· | ···················· | ···················· |
| G | 8088 hCoV-19/Ireland/CO-NVRL-72IRL13552/2020\|EPI_ISL_52 | ···················· | ·A·················· | ···················· | ···················· | ···················· | ··A················· |
| G | 8089 hCoV-19/Ireland/WD-NVRL-72IRL14394/2020\|EPI_ISL_52 | ···················· | ·A·················· | ···················· | ···················· | ···················· | ···················· |
| G | 8091 hCoV-19/Ireland/OY-NVRL-72IRL16829/2020\|EPI_ISL_52 | ···················· | ···················· | ···················· | ···················· | ···················· | ···················· |
| G | 8092 hCoV-19/Ireland/KE-NVRL-72IRL19153/2020\|EPI_ISL_52 | ···················· | ···················· | ···················· | ···················· | ···················· | ···················· |
| G | 8093 hCoV-19/Ireland/CO-NVRL-72IRL19451/2020\|EPI_ISL_52 | ···················· | ···················· | ···················· | ···················· | ···················· | ···················· |
| G | 8094 hCoV-19/Ireland/CO-NVRL-72IRL19573/2020\|EPI_ISL_52 | ···················· | ·A·················· | ···················· | ···················· | ···················· | ···················· |
| G | 8095 hCoV-19/Ireland/MH-NVRL-72IRL21078/2020\|EPI_ISL_52 | ···················· | ···················· | ···················· | ···················· | ···················· | ···················· |
| G | 8096 hCoV-19/Ireland/KK-NVRL-72IRL21404/2020\|EPI_ISL_52 | ···················· | ···················· | ···················· | ···················· | ···················· | ···················· |
| G | 8097 hCoV-19/Ireland/D-NVRL-72IRL22409/2020\|EPI_ISL_528 | ···················· | ···················· | ···················· | ···················· | ···················· | ···················· |
| G | 8098 hCoV-19/USA/AK-PHL1005/2020\|EPI_ISL_528486\|2020-07 | ···················· | ···················· | ···················· | ···················· | ···················· | ···················· |
| G | 8099 hCoV-19/USA/AK-PHL1252/2020\|EPI_ISL_528493\|2020-07 | ···················· | ···················· | ···················· | ···················· | ···················· | ···················· |
| G | 8100 hCoV-19/USA/AK-PHL1418/2020\|EPI_ISL_528494\|2020-07 | ···················· | ···················· | ···················· | ···················· | ···················· | ···················· |
| G | 8101 hCoV-19/USA/AK-PHL1420/2020\|EPI_ISL_528495\|2020-07 | ···················· | ···················· | ···················· | ···················· | ···················· | ···················· |
| G | 8102 hCoV-19/USA/AK-PHL1717/2020\|EPI_ISL_528498\|2020-07 | ···················· | ···················· | ···················· | ···················· | ···················· | ···················· |
| G | 8103 hCoV-19/Wales/PHWC-169CB5/2020\|EPI_ISL_529700\|2020 | ···················· | ···················· | ···················· | ···················· | ···················· | ···················· |
| G | 8104 hCoV-19/Wales/PHWC-169CC4/2020\|EPI_ISL_529701\|2020 | ···················· | ···················· | ···················· | ···················· | ···················· | ···················· |
| G | 8105 hCoV-19/Wales/PHWC-169CD3/2020\|EPI_ISL_529702\|2020 | ···················· | ···················· | ···················· | ···················· | ···················· | ···················· |
| G | 8106 hCoV-19/Wales/PHWC-169CE2/2020\|EPI_ISL_529703\|2020 | ···················· | ···················· | ···················· | ···················· | ···················· | ···················· |
| G | 8107 hCoV-19/Wales/PHWC-169CF1/2020\|EPI_ISL_529704\|2020 | ···················· | ···················· | ···················· | ···················· | ···················· | ···················· |
| G | 8108 hCoV-19/Wales/PHWC-169E28/2020\|EPI_ISL_529705\|2020 | ···················· | ···················· | ···················· | ···················· | ···················· | ···················· |
| G | 8109 hCoV-19/Wales/PHWC-169E64/2020\|EPI_ISL_529706\|2020 | ···················· | ···················· | ···················· | ···················· | ···················· | ···················· |
| G | 8110 hCoV-19/Wales/PHWC-169E82/2020\|EPI_ISL_529707\|2020 | ···················· | ·A·················· | ···················· | ···················· | ···················· | ··A················· |
| G | 8111 hCoV-19/Wales/PHWC-169EA0/2020\|EPI_ISL_529708\|2020 | ···················· | ···················· | ···················· | ···················· | ···················· | ···················· |
| G | 8112 hCoV-19/Wales/PHWC-169EBF/2020\|EPI_ISL_529709\|2020 | ···················· | ···················· | ···················· | ···················· | ···················· | ···················· |
| G | 8113 hCoV-19/Wales/PHWC-169EDD/2020\|EPI_ISL_529710\|2020 | ···················· | ·A·················· | ···················· | ···················· | ···················· | ··A················· |
| G | 8115 hCoV-19/Wales/PHWC-169F25/2020\|EPI_ISL_529712\|2020 | ···················· | ···················· | ···················· | ···················· | ···················· | ···················· |
| G | 8116 hCoV-19/Wales/PHWC-169F52/2020\|EPI_ISL_529713\|2020 | ···················· | ···················· | ···················· | ···················· | ···················· | ···················· |
| G | 8117 hCoV-19/Wales/PHWC-169F61/2020\|EPI_ISL_529714\|2020 | ···················· | ···················· | ···················· | ···················· | ···················· | ···················· |

| | | | | | | | |
|---|---|---|---|---|---|---|---|
| G | 8119 hCoV-19/Wales/PHWC-169F8F/2020\|EPI_ISL_529716\|2020 | .................... | .................... | .................... | .................... | .................... | .................... |
| G | 8120 hCoV-19/Wales/PHWC-169FAD/2020\|EPI_ISL_529717\|2020 | .................... | .................... | .................... | .................... | .................... | .................... |
| G | 8121 hCoV-19/South Africa/KRISP-K002717/2020\|EPI_ISL_52 | .................... | .................... | .................... | .................... | .................... | .................... |
| G | 8122 hCoV-19/South Africa/KRISP-K002720/2020\|EPI_ISL_52 | .................... | .................... | .................... | .................... | .................... | .................... |
| G | 8123 hCoV-19/South Africa/KRISP-K002722/2020\|EPI_ISL_52 | .................... | .................... | .................... | .................... | .................... | .................... |
| G | 8124 hCoV-19/South Africa/KRISP-K002724/2020\|EPI_ISL_52 | .................... | .................... | .................... | .................... | .................... | .................... |
| G | 8125 hCoV-19/South Africa/KRISP-K002727/2020\|EPI_ISL_52 | .................... | .................... | .................... | .................... | .................... | .................... |
| G | 8126 hCoV-19/South Africa/KRISP-K002729/2020\|EPI_ISL_52 | .................... | .................... | .................... | .................... | .................... | .................... |
| G | 8127 hCoV-19/South Africa/KRISP-K002731/2020\|EPI_ISL_52 | .................... | .................... | .................... | .................... | .................... | .................... |
| G | 8128 hCoV-19/South Africa/KRISP-K002732/2020\|EPI_ISL_52 | .................... | .................... | .................... | .................... | .................... | .................... |
| G | 8129 hCoV-19/South Africa/KRISP-K002733/2020\|EPI_ISL_52 | .................... | .................... | .................... | .................... | .................... | .................... |
| G | 8130 hCoV-19/South Africa/KRISP-K002734/2020\|EPI_ISL_52 | .................... | .................... | .................... | .................... | .................... | .................... |
| G | 8131 hCoV-19/South Africa/KRISP-K002735/2020\|EPI_ISL_52 | .................... | .................... | .................... | .................... | .................... | .................... |
| G | 8132 hCoV-19/South Africa/KRISP-K002736/2020\|EPI_ISL_52 | .................... | .................... | .................... | .................... | .................... | .................... |
| G | 8133 hCoV-19/South Africa/KRISP-K002737/2020\|EPI_ISL_52 | .................... | .................... | .................... | .................... | .................... | .................... |
| G | 8134 hCoV-19/South Africa/KRISP-K002738/2020\|EPI_ISL_52 | .................... | .................... | .................... | .................... | .................... | .................... |
| G | 8135 hCoV-19/South Africa/KRISP-K002739/2020\|EPI_ISL_52 | .................... | ......A............. | .................... | .................... | .................... | .................... |
| G | 8136 hCoV-19/South Africa/KRISP-K002740/2020\|EPI_ISL_52 | .................... | .................... | .................... | .................... | .................... | .................... |
| G | 8137 hCoV-19/South Africa/KRISP-K002741/2020\|EPI_ISL_52 | .................... | .................... | .................... | .................... | .................... | .................... |
| G | 8138 hCoV-19/South Africa/KRISP-K002744/2020\|EPI_ISL_52 | .................... | .................... | .................... | .................... | .................... | .................... |
| G | 8139 hCoV-19/South Africa/KRISP-K002745/2020\|EPI_ISL_52 | .................... | .................... | .................... | .................... | .................... | .................... |
| G | 8140 hCoV-19/South Africa/KRISP-K002746/2020\|EPI_ISL_52 | .................... | .................... | .................... | .................... | .................... | .................... |
| G | 8141 hCoV-19/South Africa/KRISP-K002747/2020\|EPI_ISL_52 | .................... | .................... | .................... | .................... | .................... | .................... |
| G | 8142 hCoV-19/South Africa/KRISP-K002748/2020\|EPI_ISL_52 | .................... | .................... | .................... | .................... | .................... | .................... |
| G | 8143 hCoV-19/South Africa/KRISP-K002749/2020\|EPI_ISL_52 | .................... | .................... | .................... | .................... | .................... | .................... |
| G | 8144 hCoV-19/South Africa/KRISP-K002751/2020\|EPI_ISL_52 | .................... | .................... | .................... | .................... | .................... | .................... |
| G | 8145 hCoV-19/South Africa/KRISP-K002752/2020\|EPI_ISL_52 | .................... | .................... | .................... | .................... | .................... | .................... |
| G | 8146 hCoV-19/South Africa/KRISP-K002753/2020\|EPI_ISL_52 | .................... | .................... | .................... | .................... | .................... | .................... |
| G | 8147 hCoV-19/South Africa/KRISP-K002754/2020\|EPI_ISL_52 | .................... | .................... | .................... | .................... | .................... | .................... |
| G | 8148 hCoV-19/South Africa/KRISP-K002755/2020\|EPI_ISL_52 | .................... | .................... | .................... | .................... | .................... | .................... |
| G | 8149 hCoV-19/South Africa/KRISP-K002756/2020\|EPI_ISL_52 | .................... | .................... | .................... | .................... | .................... | .................... |
| G | 8150 hCoV-19/South Africa/KRISP-K002757/2020\|EPI_ISL_52 | .................... | .................... | .................... | .................... | .................... | .................... |
| G | 8151 hCoV-19/South Africa/KRISP-K002759/2020\|EPI_ISL_52 | .................... | .................... | .................... | .................... | .................... | .................... |
| G | 8152 hCoV-19/South Africa/KRISP-K002760/2020\|EPI_ISL_52 | .................... | .................... | .................... | .................... | .................... | .................... |
| G | 8153 hCoV-19/South Africa/KRISP-K002761/2020\|EPI_ISL_52 | .................... | .................... | .................... | .................... | .................... | .................... |
| G | 8154 hCoV-19/South Africa/KRISP-K002762/2020\|EPI_ISL_52 | .................... | .................... | .................... | .................... | .................... | .................... |
| G | 8155 hCoV-19/South Africa/KRISP-K002763/2020\|EPI_ISL_52 | .................... | .................... | .................... | .................... | .................... | .................... |
| G | 8156 hCoV-19/South Africa/KRISP-K002764/2020\|EPI_ISL_52 | .................... | .................... | .................... | .................... | .................... | .................... |
| G | 8157 hCoV-19/South Africa/KRISP-K002765/2020\|EPI_ISL_52 | .................... | .................... | .................... | .................... | .................... | .................... |
| G | 8158 hCoV-19/South Africa/KRISP-K002766/2020\|EPI_ISL_52 | .................... | .................... | .................... | .................... | .................... | .................... |
| G | 8159 hCoV-19/South Africa/KRISP-K002768/2020\|EPI_ISL_52 | .................... | .................... | .................... | .................... | .................... | .................... |
| G | 8160 hCoV-19/South Africa/KRISP-K002769/2020\|EPI_ISL_52 | .................... | .................... | .................... | .................... | .................... | .................... |
| G | 8161 hCoV-19/South Africa/KRISP-K002770/2020\|EPI_ISL_52 | .................... | .................... | .................... | .................... | .................... | .................... |
| G | 8162 hCoV-19/South Africa/KRISP-K002771/2020\|EPI_ISL_52 | .................... | .................... | .................... | .................... | .................... | .................... |
| G | 8163 hCoV-19/South Africa/KRISP-K002772/2020\|EPI_ISL_52 | .................... | .................... | .................... | .................... | .................... | .................... |
| G | 8164 hCoV-19/South Africa/KRISP-K002774/2020\|EPI_ISL_52 | .................... | .................... | .................... | .................... | .................... | .................... |
| G | 8165 hCoV-19/South Africa/KRISP-K002775/2020\|EPI_ISL_52 | .................... | .................... | .................... | .................... | .................... | .................... |
| G | 8166 hCoV-19/South Africa/KRISP-K002776/2020\|EPI_ISL_52 | .................... | .................... | .................... | .................... | .................... | .................... |
| G | 8167 hCoV-19/South Africa/KRISP-K002777/2020\|EPI_ISL_52 | .................... | .................... | .................... | .................... | .................... | .................... |
| G | 8168 hCoV-19/South Africa/KRISP-K002779/2020\|EPI_ISL_52 | .................... | .................... | .................... | .................... | .................... | .................... |
| G | 8169 hCoV-19/South Africa/KRISP-K002780/2020\|EPI_ISL_52 | .................... | .................... | .................... | .................... | .................... | .................... |
| G | 8170 hCoV-19/South Africa/KRISP-K002781/2020\|EPI_ISL_52 | .................... | .................... | .................... | .................... | .................... | .................... |
| G | 8171 hCoV-19/South Africa/KRISP-K002782/2020\|EPI_ISL_52 | .................... | .................... | .................... | .................... | .................... | .................... |
| G | 8172 hCoV-19/South Africa/KRISP-K002783/2020\|EPI_ISL_52 | .................... | .................... | .................... | .................... | .................... | .................... |
| G | 8173 hCoV-19/South Africa/KRISP-K002784/2020\|EPI_ISL_52 | .................... | .................... | .................... | .................... | .................... | .................... |
| G | 8174 hCoV-19/South Africa/KRISP-K002785/2020\|EPI_ISL_52 | .................... | .................... | .................... | .................... | .................... | .................... |
| G | 8175 hCoV-19/South Africa/KRISP-K002786/2020\|EPI_ISL_52 | .................... | .................... | .................... | .................... | .................... | .................... |
| G | 8176 hCoV-19/South Africa/KRISP-K002788/2020\|EPI_ISL_52 | .................... | .................... | .................... | .................... | .................... | .................... |
| G | 8177 hCoV-19/South Africa/KRISP-K002789/2020\|EPI_ISL_52 | .................... | .................... | .................... | .................... | .................... | .................... |
| G | 8178 hCoV-19/South Africa/KRISP-K002790/2020\|EPI_ISL_52 | .................... | .................... | .................... | .................... | .................... | .................... |
| G | 8179 hCoV-19/South Africa/KRISP-K002791/2020\|EPI_ISL_52 | .................... | .................... | .................... | .................... | .................... | .................... |
| G | 8180 hCoV-19/South Africa/KRISP-K002792/2020\|EPI_ISL_52 | .................... | .................... | .................... | .................... | .................... | .................... |
| G | 8181 hCoV-19/South Africa/KRISP-K002793/2020\|EPI_ISL_52 | .................... | .................... | .................... | .................... | .................... | .................... |
| G | 8182 hCoV-19/South Africa/KRISP-K002794/2020\|EPI_ISL_52 | .................... | .................... | .................... | .................... | .................... | .................... |
| G | 8183 hCoV-19/South Africa/KRISP-K002795/2020\|EPI_ISL_52 | .................... | .................... | .................... | .................... | .................... | .................... |
| G | 8184 hCoV-19/South Africa/KRISP-K002796/2020\|EPI_ISL_52 | .................... | .................... | .................... | .................... | .................... | .................... |
| G | 8185 hCoV-19/South Africa/KRISP-K002797/2020\|EPI_ISL_52 | .................... | .................... | .................... | .................... | .................... | .................... |
| G | 8186 hCoV-19/South Africa/KRISP-K002798/2020\|EPI_ISL_52 | .................... | .................... | .................... | .................... | .................... | .................... |
| G | 8187 hCoV-19/South Africa/KRISP-K002799/2020\|EPI_ISL_52 | .................... | .................... | .................... | .................... | .................... | .................... |
| G | 8188 hCoV-19/South Africa/KRISP-K002800/2020\|EPI_ISL_52 | .................... | .................... | .................... | .................... | .................... | .................... |
| G | 8189 hCoV-19/South Africa/KRISP-K002802/2020\|EPI_ISL_52 | .................... | .................... | .................... | .................... | .................... | .................... |
| G | 8190 hCoV-19/South Africa/KRISP-K002803/2020\|EPI_ISL_52 | .................... | .................... | .................... | .................... | .................... | .................... |
| G | 8191 hCoV-19/South Africa/KRISP-K002804/2020\|EPI_ISL_52 | .................... | .................... | .................... | .................... | .................... | .................... |
| G | 8192 hCoV-19/South Africa/KRISP-K002805/2020\|EPI_ISL_52 | .................... | .................... | .................... | .................... | .................... | .................... |
| G | 8193 hCoV-19/USA/MN-MDH-1507/2020\|EPI_ISL_518900\|2020-0 | .................... | .................... | .................... | .................... | .................... | .................... |
| G | 8194 hCoV-19/Australia/VIC4490/2020\|EPI_ISL_518901\|2020 | .................... | .................... | .................... | .................... | .................... | .................... |
| G | 8195 hCoV-19/Australia/VIC4491/2020\|EPI_ISL_518902\|2020 | .................... | .................... | .................... | .................... | .................... | .................... |
| G | 8196 hCoV-19/Australia/VIC4492/2020\|EPI_ISL_518903\|2020 | .................... | .................... | .................... | .................... | .................... | .................... |
| G | 8197 hCoV-19/Australia/VIC4493/2020\|EPI_ISL_518904\|2020 | .................... | .................... | .................... | .................... | .................... | .................... |
| G | 8198 hCoV-19/Australia/VIC4494/2020\|EPI_ISL_518905\|2020 | .................... | .................... | .................... | .................... | .................... | .................... |

| | | | | | | | | |
|---|---|---|---|---|---|---|---|---|
| G | 8199 hCoV-19/Australia/VIC4495/2020\|EPI_ISL_518906\|2020 | ···················· | ···················· | ···················· | ···················· | ···················· | ···················· | |
| G | 8200 hCoV-19/Australia/VIC4496/2020\|EPI_ISL_518907\|2020 | ···················· | ···················· | ···················· | ···················· | ···················· | ···················· | |
| G | 8201 hCoV-19/Australia/VIC4497/2020\|EPI_ISL_518908\|2020 | ···················· | ···················· | ···················· | ···················· | ···················· | ···················· | |
| G | 8202 hCoV-19/Australia/VIC4498/2020\|EPI_ISL_518909\|2020 | ···················· | ···················· | ···················· | ···················· | ···················· | ···················· | |
| G | 8203 hCoV-19/Australia/VIC4499/2020\|EPI_ISL_518910\|2020 | ···················· | ···················· | ···················· | ···················· | ···················· | ···················· | |
| G | 8204 hCoV-19/Australia/VIC4500/2020\|EPI_ISL_518911\|2020 | ···················· | ···················· | ···················· | ···················· | ···················· | ···················· | |
| G | 8205 hCoV-19/Australia/VIC4501/2020\|EPI_ISL_518912\|2020 | ···················· | ···················· | ···················· | ···················· | ···················· | ···················· | |
| G | 8206 hCoV-19/Australia/VIC4502/2020\|EPI_ISL_518913\|2020 | ···················· | ···················· | ···················· | ···················· | ···················· | ···················· | |
| G | 8207 hCoV-19/Australia/VIC4503/2020\|EPI_ISL_518914\|2020 | ···················· | ···················· | ···················· | ···················· | ···················· | ···················· | |
| G | 8208 hCoV-19/Australia/VIC4504/2020\|EPI_ISL_518915\|2020 | ···················· | ···················· | ···················· | ···················· | ···················· | ···················· | |
| G | 8209 hCoV-19/Australia/VIC4506/2020\|EPI_ISL_518917\|2020 | ···················· | ···················· | ···················· | ···················· | ···················· | ···················· | |
| G | 8210 hCoV-19/Australia/VIC4508/2020\|EPI_ISL_518919\|2020 | ···················· | ···················· | ···················· | ···················· | ···················· | ···················· | |
| G | 8211 hCoV-19/Australia/VIC4509/2020\|EPI_ISL_518920\|2020 | ···················· | ···················· | ···················· | ···················· | ···················· | ···················· | |
| G | 8212 hCoV-19/Australia/VIC4510/2020\|EPI_ISL_518921\|2020 | ···················· | ···················· | ···················· | ···················· | ···················· | ···················· | |
| G | 8213 hCoV-19/Australia/VIC4511/2020\|EPI_ISL_518922\|2020 | ···················· | ···················· | ···················· | ···················· | ···················· | ···················· | |
| G | 8214 hCoV-19/Australia/VIC4512/2020\|EPI_ISL_518923\|2020 | ···················· | ···················· | ···················· | ···················· | ···················· | ···················· | |
| G | 8215 hCoV-19/Australia/VIC4513/2020\|EPI_ISL_518924\|2020 | ···················· | ···················· | ···················· | ···················· | ···················· | ···················· | |
| G | 8216 hCoV-19/Australia/VIC4514/2020\|EPI_ISL_518925\|2020 | ···················· | ···················· | ···················· | ···················· | ···················· | ···················· | |
| G | 8217 hCoV-19/Australia/VIC4515/2020\|EPI_ISL_518926\|2020 | ···················· | ···················· | ···················· | ···················· | ···················· | ···················· | |
| G | 8218 hCoV-19/Australia/VIC2388/2020\|EPI_ISL_522106\|2020 | ···················· | ···················· | ···················· | ···················· | ···················· | ···················· | |
| G | 8219 hCoV-19/Australia/VIC2389/2020\|EPI_ISL_522107\|2020 | ···················· | ···················· | ···················· | ···················· | ···················· | ···················· | |
| G | 8220 hCoV-19/Australia/VIC2390/2020\|EPI_ISL_522108\|2020 | ···················· | ·A·················· | ···················· | ···················· | ···················· | ··A················· | |
| G | 8221 hCoV-19/Australia/VIC2392/2020\|EPI_ISL_522109\|2020 | ···················· | ···················· | ···················· | ···················· | ···················· | ···················· | |
| G | 8222 hCoV-19/Australia/VIC2394/2020\|EPI_ISL_522111\|2020 | ···················· | ···················· | ···················· | ···················· | ···················· | ···················· | |
| G | 8224 hCoV-19/Australia/VIC2397/2020\|EPI_ISL_522113\|2020 | ···················· | ···················· | ···················· | ···················· | ···················· | ···················· | |
| G | 8225 hCoV-19/Australia/VIC2398/2020\|EPI_ISL_522114\|2020 | ···················· | ···················· | ···················· | ···················· | ···················· | ···················· | |
| G | 8226 hCoV-19/Australia/VIC2399/2020\|EPI_ISL_522115\|2020 | ···················· | ···················· | ···················· | ···················· | ···················· | ···················· | |
| G | 8227 hCoV-19/Australia/VIC2400/2020\|EPI_ISL_522116\|2020 | ···················· | ···················· | ···················· | ···················· | ···················· | ···················· | |
| G | 8229 hCoV-19/Australia/VIC2402/2020\|EPI_ISL_522118\|2020 | ···················· | ···················· | ···················· | ···················· | ···················· | ···················· | |
| G | 8230 hCoV-19/Australia/VIC2409/2020\|EPI_ISL_522120\|2020 | ···················· | ···················· | ···················· | ···················· | ···················· | ···················· | |
| G | 8231 hCoV-19/Australia/VIC2410/2020\|EPI_ISL_522121\|2020 | ···················· | ···················· | ···················· | ···················· | ···················· | ···················· | |
| G | 8232 hCoV-19/Australia/VIC2411/2020\|EPI_ISL_522122\|2020 | ···················· | ···················· | ···················· | ···················· | ···················· | ···················· | |
| G | 8233 hCoV-19/Australia/VIC2412/2020\|EPI_ISL_522123\|2020 | ···················· | ···················· | ···················· | ···················· | ···················· | ···················· | |
| G | 8234 hCoV-19/Australia/VIC2413/2020\|EPI_ISL_522124\|2020 | ···················· | ···················· | ···················· | ···················· | ···················· | ···················· | |
| G | 8235 hCoV-19/Australia/VIC2414/2020\|EPI_ISL_522125\|2020 | ···················· | ···················· | ···················· | ···················· | ···················· | ···················· | |
| G | 8236 hCoV-19/Australia/VIC2415/2020\|EPI_ISL_522126\|2020 | ···················· | ···················· | ···················· | ···················· | ···················· | ···················· | |
| G | 8237 hCoV-19/Australia/VIC2416/2020\|EPI_ISL_522127\|2020 | ···················· | ···················· | ···················· | ···················· | ···················· | ···················· | |
| G | 8238 hCoV-19/Australia/VIC2417/2020\|EPI_ISL_522128\|2020 | ···················· | ···················· | ···················· | ···················· | ···················· | ···················· | |
| G | 8239 hCoV-19/Australia/VIC2418/2020\|EPI_ISL_522129\|2020 | ···················· | ···················· | ···················· | ···················· | ···················· | ···················· | |
| G | 8240 hCoV-19/Australia/VIC2420/2020\|EPI_ISL_522130\|2020 | ···················· | ···················· | ···················· | ···················· | ···················· | ···················· | |
| G | 8241 hCoV-19/Australia/VIC2421/2020\|EPI_ISL_522131\|2020 | ···················· | ···················· | ···················· | ···················· | ···················· | ···················· | |
| G | 8242 hCoV-19/Australia/VIC2422/2020\|EPI_ISL_522132\|2020 | ···················· | ···················· | ···················· | ···················· | ···················· | ···················· | |
| G | 8243 hCoV-19/Australia/VIC2423/2020\|EPI_ISL_522133\|2020 | ···················· | ···················· | ···················· | ···················· | ···················· | ···················· | |
| G | 8244 hCoV-19/Australia/VIC2424/2020\|EPI_ISL_522134\|2020 | ···················· | ···················· | ···················· | ···················· | ···················· | ···················· | |
| G | 8245 hCoV-19/Australia/VIC2425/2020\|EPI_ISL_522135\|2020 | ···················· | ···················· | ···················· | ···················· | ···················· | ···················· | |
| G | 8246 hCoV-19/Australia/VIC2426/2020\|EPI_ISL_522136\|2020 | ···················· | ···················· | ···················· | ···················· | NNN················· | ···················· | 1 |
| G | 8247 hCoV-19/Australia/VIC2427/2020\|EPI_ISL_522137\|2020 | ···················· | ···················· | ···················· | ···················· | ···················· | ···················· | |
| G | 8248 hCoV-19/Australia/VIC2429/2020\|EPI_ISL_522138\|2020 | ···················· | ···················· | ···················· | ···················· | ···················· | ···················· | |
| G | 8249 hCoV-19/Australia/VIC2430/2020\|EPI_ISL_522139\|2020 | ···················· | ···················· | ···················· | ···················· | ···················· | ···················· | |
| G | 8250 hCoV-19/Australia/VIC2431/2020\|EPI_ISL_522140\|2020 | ···················· | ···················· | ···················· | ···················· | ···················· | ···················· | |
| G | 8251 hCoV-19/Australia/VIC2432/2020\|EPI_ISL_522141\|2020 | ···················· | ···················· | ···················· | ···················· | ···················· | ···················· | |
| G | 8252 hCoV-19/Australia/VIC2433/2020\|EPI_ISL_522142\|2020 | ···················· | ···················· | ···················· | ···················· | ···················· | ···················· | |
| G | 8253 hCoV-19/Australia/VIC2435/2020\|EPI_ISL_522143\|2020 | ···················· | ···················· | ···················· | ···················· | ···················· | ···················· | |
| G | 8254 hCoV-19/Australia/VIC2437/2020\|EPI_ISL_522144\|2020 | ···················· | ···················· | ···················· | ···················· | ·NN················· | ···················· | |
| G | 8255 hCoV-19/Australia/VIC2439/2020\|EPI_ISL_522145\|2020 | ···················· | ···················· | ···················· | ···················· | ···················· | ···················· | |
| G | 8256 hCoV-19/Australia/VIC2440/2020\|EPI_ISL_522146\|2020 | ···················· | ···················· | ···················· | ···················· | ···················· | ···················· | |
| G | 8257 hCoV-19/Australia/VIC2441/2020\|EPI_ISL_522147\|2020 | ···················· | ···················· | ···················· | ···················· | ···················· | ···················· | |
| G | 8258 hCoV-19/Australia/VIC2442/2020\|EPI_ISL_522148\|2020 | ···················· | ···················· | ···················· | ···················· | ···················· | ···················· | |
| G | 8259 hCoV-19/Australia/VIC2443/2020\|EPI_ISL_522149\|2020 | ···················· | ···················· | ···················· | ···················· | ···················· | ···················· | |
| G | 8260 hCoV-19/Australia/VIC2444/2020\|EPI_ISL_522150\|2020 | ···················· | ···················· | ···················· | ···················· | ···················· | ···················· | |
| G | 8261 hCoV-19/Australia/VIC2445/2020\|EPI_ISL_522151\|2020 | ···················· | ···················· | ···················· | ···················· | ···················· | ···················· | |
| G | 8262 hCoV-19/Australia/VIC2446/2020\|EPI_ISL_522152\|2020 | ···················· | ···················· | ···················· | ···················· | ···················· | ···················· | |
| G | 8263 hCoV-19/Australia/VIC2447/2020\|EPI_ISL_522153\|2020 | ···················· | ···················· | ···················· | ···················· | ···················· | ···················· | |
| G | 8264 hCoV-19/Australia/VIC2448/2020\|EPI_ISL_522154\|2020 | ···················· | ···················· | ···················· | ···················· | ···················· | ···················· | |
| G | 8265 hCoV-19/Australia/VIC2449/2020\|EPI_ISL_522155\|2020 | ···················· | ···················· | ···················· | ···················· | ···················· | ···················· | |
| G | 8266 hCoV-19/Australia/VIC2451/2020\|EPI_ISL_522157\|2020 | ···················· | ···················· | ···················· | ···················· | ···················· | ···················· | |
| G | 8267 hCoV-19/Australia/VIC2452/2020\|EPI_ISL_522158\|2020 | ···················· | ···················· | ···················· | ···················· | ···················· | ···················· | |
| G | 8268 hCoV-19/Australia/VIC2453/2020\|EPI_ISL_522159\|2020 | ···················· | ···················· | ···················· | ···················· | ···················· | ···················· | |
| G | 8269 hCoV-19/Australia/VIC2454/2020\|EPI_ISL_522160\|2020 | ···················· | ···················· | ···················· | ···················· | ···················· | ···················· | |
| G | 8270 hCoV-19/Australia/VIC2455/2020\|EPI_ISL_522161\|2020 | ···················· | ···················· | ···················· | ···················· | ···················· | ···················· | |
| G | 8271 hCoV-19/Australia/VIC2456/2020\|EPI_ISL_522162\|2020 | ···················· | ···················· | ···················· | ···················· | ···················· | ···················· | |
| G | 8272 hCoV-19/Australia/VIC2457/2020\|EPI_ISL_522163\|2020 | ···················· | ···················· | ···················· | ···················· | ···················· | ···················· | |
| G | 8273 hCoV-19/Australia/VIC2458/2020\|EPI_ISL_522164\|2020 | ···················· | ···················· | ···················· | ···················· | ···················· | ···················· | |
| G | 8274 hCoV-19/Australia/VIC2459/2020\|EPI_ISL_522165\|2020 | ···················· | ···················· | ···················· | ···················· | ···················· | ···················· | |
| G | 8275 hCoV-19/Australia/VIC2460/2020\|EPI_ISL_522166\|2020 | ···················· | ···················· | ···················· | ···················· | ···················· | ···················· | |
| G | 8276 hCoV-19/Australia/VIC2461/2020\|EPI_ISL_522167\|2020 | ···················· | ···················· | ···················· | ···················· | ···················· | ···················· | |
| G | 8277 hCoV-19/Australia/VIC2462/2020\|EPI_ISL_522168\|2020 | ···················· | ···················· | ···················· | ···················· | ···················· | ···················· | |
| G | 8278 hCoV-19/Australia/VIC2463/2020\|EPI_ISL_522169\|2020 | ···················· | ···················· | ···················· | ···················· | ···················· | ···················· | |
| G | 8279 hCoV-19/Australia/VIC2464/2020\|EPI_ISL_522170\|2020 | ···················· | ···················· | ···················· | ···················· | ···················· | ···················· | |
| G | 8280 hCoV-19/Australia/VIC2468/2020\|EPI_ISL_522171\|2020 | ···················· | ···················· | ··A················· | ···················· | ···················· | ···················· | |

G 8281 hCoV-19/Australia/VIC2469/2020|EPI_ISL_522172|2020 ···················· ···················· ···················· ···················· ···················· ····················
G 8282 hCoV-19/Australia/VIC2470/2020|EPI_ISL_522173|2020 ···················· ···················· ···················· ···················· ···················· ····················
G 8283 hCoV-19/Australia/VIC2471/2020|EPI_ISL_522174|2020 ···················· ···················· ···················· ···················· ···················· ····················
G 8284 hCoV-19/Australia/VIC2472/2020|EPI_ISL_522175|2020 ···················· ···················· ···················· ···················· ···················· ····················
G 8285 hCoV-19/Australia/VIC2473/2020|EPI_ISL_522176|2020 ···················· ···················· ···················· ···················· ···················· ····················
G 8286 hCoV-19/Australia/VIC2474/2020|EPI_ISL_522177|2020 ···················· ···················· ···················· ···················· ···················· ····················
G 8287 hCoV-19/Australia/VIC2475/2020|EPI_ISL_522178|2020 ···················· ···················· ···················· ···················· ···················· ····················
G 8288 hCoV-19/Australia/VIC2476/2020|EPI_ISL_522179|2020 ···················· ···················· ···················· ···················· ···················· ····················
G 8289 hCoV-19/Australia/VIC2477/2020|EPI_ISL_522180|2020 ···················· ···················· ···················· ···················· ···················· ····················
G 8290 hCoV-19/Australia/VIC2478/2020|EPI_ISL_522181|2020 ···················· ···················· ···················· ···················· ···················· ····················
G 8291 hCoV-19/Australia/VIC2479/2020|EPI_ISL_522182|2020 ···················· ···················· ···················· ···················· ···················· ····················
G 8292 hCoV-19/Australia/VIC2480/2020|EPI_ISL_522183|2020 ···················· ···················· ···················· ···················· ···················· ····················
G 8293 hCoV-19/Australia/VIC2481/2020|EPI_ISL_522184|2020 ···················· ···················· ···················· ···················· ···················· ····················
G 8295 hCoV-19/Australia/VIC2484/2020|EPI_ISL_522186|2020 ···················· ···················· ···················· ···················· ···················· ····················
G 8296 hCoV-19/Australia/VIC2489/2020|EPI_ISL_522187|2020 ···················· ···················· ···················· ···················· ···················· ····················
G 8297 hCoV-19/Australia/VIC2490/2020|EPI_ISL_522188|2020 ···················· ···················· ···················· ···················· ···················· ····················
G 8298 hCoV-19/Australia/VIC2491/2020|EPI_ISL_522189|2020 ···················· ···················· ···················· ···················· ···················· ····················
G 8299 hCoV-19/Netherlands/NB-EMC-279/2020|EPI_ISL_523401 ···················· ···················· ···················· ···················· ···················· ····················
G 8300 hCoV-19/Netherlands/NB-EMC-280/2020|EPI_ISL_523402 ···················· ···················· ···················· ···················· ···················· ····················
G 8301 hCoV-19/Netherlands/NB-EMC-281/2020|EPI_ISL_523403 ···················· ···················· ···················· ···················· ···················· ····················
G 8302 hCoV-19/Netherlands/NH-EMC-36/2020|EPI_ISL_523405| ···················· ···················· ···················· ···················· ···················· ····················
G 8303 hCoV-19/Netherlands/NH-EMC-38/2020|EPI_ISL_523407| ···················· ···················· ···················· ···················· ···················· ····················
G 8304 hCoV-19/Netherlands/NH-EMC-39/2020|EPI_ISL_523408| ···················· ···················· ···················· ···················· ···················· ····················
G 8305 hCoV-19/Netherlands/NH-EMC-40/2020|EPI_ISL_523409| ···················· ···················· ···················· ···················· ···················· ····················
G 8306 hCoV-19/Netherlands/NH-EMC-43/2020|EPI_ISL_523412| ···················· ···················· ···················· ···················· ···················· ····················
G 8307 hCoV-19/Netherlands/NH-EMC-44/2020|EPI_ISL_523413| ···················· ···················· ···················· ···················· ···················· ····················
G 8308 hCoV-19/Netherlands/NH-EMC-45/2020|EPI_ISL_523414| ···················· ···················· ···················· ···················· ···················· ····················
G 8309 hCoV-19/Netherlands/OV-EMC-53/2020|EPI_ISL_523437| ···················· ···········A········ ···················· ···················· ···················· ····················
G 8310 hCoV-19/Netherlands/OV-EMC-55/2020|EPI_ISL_523439| ···················· ···················· ···················· ···················· ···················· ····················
G 8311 hCoV-19/Netherlands/OV-EMC-57/2020|EPI_ISL_523441| ···················· ···················· ···················· ···················· ···················· ····················
G 8312 hCoV-19/Netherlands/OV-EMC-58/2020|EPI_ISL_523442| ···················· ···················· ···················· ···················· ···················· ····················
G 8313 hCoV-19/Netherlands/OV-EMC-59/2020|EPI_ISL_523443| ···················· ···················· ···················· ···················· ···················· ····················
G 8314 hCoV-19/Netherlands/OV-EMC-60/2020|EPI_ISL_523444| ···················· ···················· ···················· ···················· ···················· ····················
G 8315 hCoV-19/Netherlands/OV-EMC-61/2020|EPI_ISL_523445| ···················· ···················· ···················· ···················· ···················· ····················
G 8316 hCoV-19/Netherlands/OV-EMC-62/2020|EPI_ISL_523446| ···················· ···················· ···················· ···················· ···················· ····················
G 8317 hCoV-19/Netherlands/OV-EMC-63/2020|EPI_ISL_523447| ···················· ···················· ···················· ···················· ···················· ····················
G 8318 hCoV-19/Netherlands/OV-EMC-64/2020|EPI_ISL_523448| ···················· ···················· ···················· ···················· ···················· ····················
G 8319 hCoV-19/Netherlands/OV-EMC-65/2020|EPI_ISL_523449| ···················· ···················· ···················· ···················· ···················· ····················
G 8320 hCoV-19/Netherlands/OV-EMC-66/2020|EPI_ISL_523450| ···················· ···················· ···················· ···················· ···················· ····················
G 8321 hCoV-19/Netherlands/OV-EMC-67/2020|EPI_ISL_523451| ···················· ···················· ···················· ···················· ···················· ····················
G 8322 hCoV-19/Netherlands/OV-EMC-68/2020|EPI_ISL_523452| ···················· ···················· ···················· ···················· ···················· ····················
G 8323 hCoV-19/Netherlands/OV-EMC-69/2020|EPI_ISL_523453| ···················· ···················· ···················· ···················· ···················· ····················
G 8324 hCoV-19/Netherlands/OV-EMC-70/2020|EPI_ISL_523454| ···················· ···················· ···················· ···················· ···················· ····················
G 8325 hCoV-19/Netherlands/OV-EMC-71/2020|EPI_ISL_523455| ···················· ···················· ···················· ···················· ···················· ····················
G 8326 hCoV-19/Netherlands/OV-EMC-72/2020|EPI_ISL_523456| ···················· ···················· ···················· ···················· ···················· ····················
G 8327 hCoV-19/Netherlands/OV-EMC-73/2020|EPI_ISL_523457| ···················· ···················· ···················· ···················· ···················· ····················
G 8328 hCoV-19/Netherlands/OV-EMC-74/2020|EPI_ISL_523458| ···················· ···················· ···················· ···················· ···················· ····················
G 8329 hCoV-19/Netherlands/OV-EMC-75/2020|EPI_ISL_523459| ···················· ···················· ···················· ···················· ···················· ····················
G 8330 hCoV-19/Netherlands/OV-EMC-76/2020|EPI_ISL_523460| ···················· ·A·················· ···················· ···················· ···················· ····················
G 8331 hCoV-19/Netherlands/OV-EMC-77/2020|EPI_ISL_523461| ···················· ···················· ···················· ···················· ···················· ····················
G 8332 hCoV-19/Netherlands/OV-EMC-78/2020|EPI_ISL_523462| ···················· ···················· ···················· ···················· ···················· ····················
G 8333 hCoV-19/Netherlands/OV-EMC-79/2020|EPI_ISL_523463| ···················· ···················· ···················· ···················· ···················· ····················
G 8334 hCoV-19/Netherlands/OV-EMC-80/2020|EPI_ISL_523464| ···················· ···················· ···················· ···················· ···················· ····················
G 8335 hCoV-19/Netherlands/OV-EMC-81/2020|EPI_ISL_523465| ···················· ···················· ···················· ···················· ···················· ····················
G 8336 hCoV-19/Netherlands/un-EMC-711/2020|EPI_ISL_523466 ···················· ···················· ···················· ···················· ···················· ····················
G 8337 hCoV-19/Netherlands/un-EMC-712/2020|EPI_ISL_523467 ···················· ···················· ···················· ···················· ···················· ····················
G 8338 hCoV-19/Netherlands/un-EMC-713/2020|EPI_ISL_523468 ···················· ···················· ···················· ···················· ···················· ····················
G 8339 hCoV-19/Netherlands/un-EMC-714/2020|EPI_ISL_523469 ···················· ···················· ···················· ···················· ···················· ····················
G 8340 hCoV-19/Netherlands/un-EMC-715/2020|EPI_ISL_523470 ···················· ···················· ···················· ···················· ···················· ····················
G 8341 hCoV-19/Netherlands/un-EMC-716/2020|EPI_ISL_523471 ···················· ···················· ···················· ···················· ···················· ····················
G 8342 hCoV-19/Netherlands/un-EMC-717/2020|EPI_ISL_523472 ···················· ···················· ···················· ···················· ···················· ····················
G 8343 hCoV-19/Netherlands/un-EMC-718/2020|EPI_ISL_523473 ···················· ···················· ···················· ···················· ···················· ····················
G 8344 hCoV-19/Netherlands/un-EMC-719/2020|EPI_ISL_523474 ···················· ···················· ···················· ···················· ···················· ····················
G 8345 hCoV-19/Netherlands/un-EMC-720/2020|EPI_ISL_523475 ···················· ···················· ···················· ···················· ···················· ····················
G 8346 hCoV-19/Netherlands/un-EMC-721/2020|EPI_ISL_523476 ···················· ···················· ···················· ···················· ···················· ····················
G 8347 hCoV-19/Netherlands/un-EMC-724/2020|EPI_ISL_523479 ···················· ···················· ···················· ···················· ···················· ····················
G 8348 hCoV-19/Netherlands/un-EMC-728/2020|EPI_ISL_523483 ···················· ···················· ···················· ···················· ···················· ····················
G 8349 hCoV-19/Netherlands/un-EMC-729/2020|EPI_ISL_523484 ···················· ···················· ···················· ···················· ···················· ····················
G 8350 hCoV-19/Netherlands/un-EMC-730/2020|EPI_ISL_523485 ···················· ···················· ···················· ···················· ···················· ····················
G 8351 hCoV-19/Netherlands/un-EMC-731/2020|EPI_ISL_523486 ···················· ···················· ···················· ···················· ···················· ····················
G 8352 hCoV-19/Netherlands/un-EMC-732/2020|EPI_ISL_523487 ···················· ···················· ···················· ···················· ···················· ····················
G 8353 hCoV-19/Netherlands/un-EMC-735/2020|EPI_ISL_523490 ···················· ···················· ···················· ···················· ···················· ····················
G 8354 hCoV-19/Netherlands/un-EMC-736/2020|EPI_ISL_523491 ···················· ···················· ···················· ···················· ···················· ····················
G 8355 hCoV-19/Netherlands/un-EMC-738/2020|EPI_ISL_523493 ···················· ···················· ···················· ···················· ···················· ····················
G 8356 hCoV-19/Netherlands/un-EMC-739/2020|EPI_ISL_523494 ···················· ···················· ···················· ···················· ···················· ····················
G 8357 hCoV-19/Netherlands/un-EMC-740/2020|EPI_ISL_523495 ···················· ···················· ···················· ···················· ···················· ····················
G 8358 hCoV-19/Netherlands/un-EMC-742/2020|EPI_ISL_523497 ···················· ···········A········ ···················· ···················· ···················· ····················
G 8359 hCoV-19/Netherlands/un-EMC-743/2020|EPI_ISL_523498 ···················· ···················· ···················· ···················· ···················· ····················
G 8360 hCoV-19/Netherlands/un-EMC-744/2020|EPI_ISL_523499 ···················· ···················· ···················· ···················· ···················· ····················
G 8361 hCoV-19/England/LOND-12607E6/2020|EPI_ISL_524700|2 ···················· ·············M······ ···················· ···················· ···················· ····················

| | | | | | | | | | |
|---|---|---|---|---|---|---|---|---|---|
| G | 8362 | hCoV-19/England/LOND-12616CA/2020\|EPI_ISL_524702\|2 | ···················· | ···················· | ····················· | ···················· | ···················· | ···················· | |
| G | 8363 | hCoV-19/England/LOND-126028B/2020\|EPI_ISL_524703\|2 | ···················· | ···················· | ····················· | ···················· | ···················· | ···················· | |
| G | 8364 | hCoV-19/England/LOND-1260F75/2020\|EPI_ISL_524704\|2 | ···················· | ···················· | ····················· | ···················· | ···················· | ···················· | |
| G | 8365 | hCoV-19/England/LOND-1261062/2020\|EPI_ISL_524705\|2 | ···················· | ···················· | ····················· | ···················· | ···················· | ···················· | |
| G | 8366 | hCoV-19/England/LOND-126169D/2020\|EPI_ISL_524706\|2 | ···················· | ···················· | ····················· | ···················· | ···················· | ···················· | |
| G | 8367 | hCoV-19/England/LOND-1261536/2020\|EPI_ISL_524707\|2 | ···················· | ···················· | ····················· | ···················· | ···················· | ···················· | |
| G | 8368 | hCoV-19/England/LOND-1261448/2020\|EPI_ISL_524708\|2 | ···················· | ···················· | ····················· | ···················· | ···················· | ···················· | |
| G | 8369 | hCoV-19/England/LOND-1260E3C/2020\|EPI_ISL_524709\|2 | ···················· | ···················· | ····················· | ···················· | ···················· | ···················· | |
| G | 8370 | hCoV-19/England/LOND-126079B/2020\|EPI_ISL_524710\|2 | ···················· | ···················· | ····················· | ···················· | ···················· | ···················· | |
| G | 8371 | hCoV-19/England/LOND-12614DF/2020\|EPI_ISL_524711\|2 | ···················· | ···················· | ····················· | ···················· | ···················· | ···················· | |
| G | 8372 | hCoV-19/England/LOND-1261527/2020\|EPI_ISL_524712\|2 | ···················· | ···················· | ····················· | ···················· | ···················· | ···················· | |
| G | 8373 | hCoV-19/India/GJ-GBRC-362a/2020\|EPI_ISL_524713\|202 | ···················· | ···················· | ····················· | ···················· | ···················· | ···················· | |
| G | 8374 | hCoV-19/India/GJ-GBRC-362b/2020\|EPI_ISL_524714\|202 | ···················· | ···················· | ····················· | ···················· | ···················· | ···················· | |
| G | 8375 | hCoV-19/India/GJ-GBRC-363/2020\|EPI_ISL_524715\|2020 | ···················· | ·A·················· | ····················· | ···················· | ···················· | ···A················ | |
| G | 8376 | hCoV-19/India/GJ-GBRC-364a/2020\|EPI_ISL_524716\|202 | ···················· | ·A·················· | ····················· | ···················· | ···················· | ···A················ | |
| G | 8377 | hCoV-19/India/GJ-GBRC-364b/2020\|EPI_ISL_524717\|202 | ···················· | ·A·················· | ····················· | ···················· | ···················· | ···A················ | |
| G | 8378 | hCoV-19/India/GJ-GBRC-365/2020\|EPI_ISL_524718\|2020 | ···················· | ·A·················· | ····················· | ···················· | ···················· | ···A················ | |
| G | 8379 | hCoV-19/India/GJ-GBRC-366a/2020\|EPI_ISL_524719\|202 | ···················· | ····A··············· | ····················· | ···················· | ···················· | ···················· | |
| G | 8380 | hCoV-19/India/GJ-GBRC-366b/2020\|EPI_ISL_524720\|202 | ···················· | ····A··············· | ····················· | ···················· | ···················· | ···················· | |
| G | 8381 | hCoV-19/India/GJ-GBRC-367a/2020\|EPI_ISL_524721\|202 | ···················· | ·A·················· | ····················· | ···················· | ···················· | ···A················ | |
| G | 8382 | hCoV-19/India/GJ-GBRC-367b/2020\|EPI_ISL_524722\|202 | ···················· | ·A·················· | ····················· | ···················· | ···················· | ···A················ | |
| G | 8383 | hCoV-19/India/GJ-GBRC-368a/2020\|EPI_ISL_524723\|202 | ···················· | ·A·················· | ····················· | ···················· | ···················· | ···A················ | |
| G | 8384 | hCoV-19/India/GJ-GBRC-368b/2020\|EPI_ISL_524724\|202 | ···················· | ·A·················· | ····················· | ···················· | ···················· | ···A················ | |
| G | 8385 | hCoV-19/India/GJ-GBRC-369/2020\|EPI_ISL_524725\|2020 | ···················· | ···················· | ····················· | ···················· | ···················· | ···················· | |
| G | 8386 | hCoV-19/India/GJ-GBRC-370/2020\|EPI_ISL_524726\|2020 | ···················· | ·A·················· | ····················· | ···················· | ···················· | ···A················ | |
| G | 8387 | hCoV-19/India/GJ-GBRC-371/2020\|EPI_ISL_524727\|2020 | ···················· | ·A·················· | ····················· | ···················· | ···················· | ···A················ | |
| G | 8388 | hCoV-19/India/GJ-GBRC-372/2020\|EPI_ISL_524728\|2020 | ···················· | ·A·················· | ····················· | ···················· | ···················· | ···A················ | |
| G | 8389 | hCoV-19/India/GJ-GBRC-373/2020\|EPI_ISL_524729\|2020 | ···················· | ···················· | ····················· | ···················· | ···················· | ···················· | |
| G | 8390 | hCoV-19/India/GJ-GBRC-374a/2020\|EPI_ISL_524730\|202 | ···················· | ·A·················· | ····················· | ···················· | ···················· | ···A················ | |
| G | 8391 | hCoV-19/India/GJ-GBRC-374b/2020\|EPI_ISL_524731\|202 | ···················· | ·A·················· | ····················· | ···················· | ···················· | ···A················ | |
| G | 8392 | hCoV-19/India/GJ-GBRC-375/2020\|EPI_ISL_524732\|2020 | ···················· | ···················· | ····················· | ···················· | ···················· | ···················· | |
| G | 8393 | hCoV-19/India/GJ-GBRC-376a/2020\|EPI_ISL_524733\|202 | ···················· | ···················· | ····················· | ···················· | ···················· | ···················· | |
| G | 8394 | hCoV-19/India/GJ-GBRC-376b/2020\|EPI_ISL_524734\|202 | ···················· | ···················· | ····················· | ···················· | ···················· | ···················· | |
| G | 8395 | hCoV-19/India/GJ-GBRC-377a/2020\|EPI_ISL_524735\|202 | ···················· | ·A·················· | ····················· | ···················· | ···················· | ···A················ | |
| G | 8396 | hCoV-19/India/GJ-GBRC-377b/2020\|EPI_ISL_524736\|202 | ···················· | ·A·················· | ····················· | ···················· | ···················· | ···················· | |
| G | 8397 | hCoV-19/India/GJ-GBRC-378/2020\|EPI_ISL_524737\|2020 | ···················· | ·A·················· | ····················· | ···················· | ···················· | ···A················ | |
| G | 8398 | hCoV-19/India/GJ-GBRC-379/2020\|EPI_ISL_524738\|2020 | ···················· | ·A·················· | ····················· | ···················· | ···················· | ···A················ | |
| G | 8399 | hCoV-19/India/GJ-GBRC-380a/2020\|EPI_ISL_524739\|202 | ···················· | ·A·················· | ····················· | ···················· | ···················· | ···A················ | |
| G | 8400 | hCoV-19/India/GJ-GBRC-380b/2020\|EPI_ISL_524740\|202 | ···················· | ·A·················· | ····················· | ···················· | ···················· | ···A················ | |
| G | 8401 | hCoV-19/India/GJ-GBRC-381/2020\|EPI_ISL_524741\|2020 | ···················· | ·A·················· | ····················· | ···················· | ···················· | ···A················ | |
| G | 8402 | hCoV-19/India/GJ-GBRC-383b/2020\|EPI_ISL_524743\|202 | ···················· | ·A·················· | ····················· | ···················· | ···················· | ···A················ | |
| G | 8403 | hCoV-19/India/GJ-GBRC-384a/2020\|EPI_ISL_524744\|202 | ···················· | ·A·················· | ····················· | ···················· | ···················· | ···A················ | |
| G | 8404 | hCoV-19/India/GJ-GBRC-384b/2020\|EPI_ISL_524745\|202 | ···················· | ·A·················· | ····················· | ···················· | ···················· | ···················· | |
| G | 8405 | hCoV-19/USA/UT-08710/2020\|EPI_ISL_524768\|2020-05-1 | ···················· | ···················· | ····················· | ···················· | ···················· | ···················· | |
| G | 8406 | hCoV-19/USA/UT-08711/2020\|EPI_ISL_524769\|2020-05-1 | ···················· | ···················· | ····················· | ···················· | ···················· | ···················· | |
| G | 8407 | hCoV-19/USA/UT-08712/2020\|EPI_ISL_524770\|2020-05-1 | ···················· | ···················· | ····················· | ···················· | ···················· | ···················· | |
| G | 8408 | hCoV-19/USA/UT-08714/2020\|EPI_ISL_524771\|2020-05-1 | ···················· | ···················· | ····················· | ···················· | ···················· | ···················· | |
| G | 8409 | hCoV-19/USA/UT-08715/2020\|EPI_ISL_524772\|2020-05-1 | ···················· | ···················· | ····················· | ···················· | ···················· | ···················· | |
| G | 8410 | hCoV-19/USA/UT-08716/2020\|EPI_ISL_524773\|2020-05-1 | ···················· | ···················· | ····················· | ···················· | ···················· | ···················· | |
| G | 8411 | hCoV-19/USA/UT-08717/2020\|EPI_ISL_524774\|2020-05-2 | ···················· | ···················· | ····················· | ···················· | ···················· | ···················· | |
| G | 8412 | hCoV-19/USA/UT-08718/2020\|EPI_ISL_524775\|2020-05-2 | ···················· | ···················· | ····················· | ···················· | ···················· | ···················· | |
| G | 8413 | hCoV-19/USA/UT-08719/2020\|EPI_ISL_524776\|2020-05-2 | ···················· | ···················· | ····················· | ···················· | ·········A·········· | ···················· | |
| G | 8414 | hCoV-19/USA/UT-08721/2020\|EPI_ISL_524778\|2020-05-2 | ···················· | ···················· | ····················· | ···················· | ···················· | ···················· | |
| G | 8415 | hCoV-19/USA/UT-08722/2020\|EPI_ISL_524779\|2020-05-2 | ···················· | ···················· | ····················· | ···················· | ···················· | ···················· | |
| G | 8416 | hCoV-19/USA/UT-08723/2020\|EPI_ISL_524780\|2020-05-2 | ···················· | ···················· | ····················· | ···················· | ···················· | ···················· | |
| G | 8417 | hCoV-19/Switzerland/ZH-UZH-1000477909/2020\|EPI_ISL | ···················· | ···················· | ····················· | ···················· | ···················· | ···················· | |
| G | 8418 | hCoV-19/Brazil/PA-IEC-164747/2020\|EPI_ISL_524783\|2 | ···················· | ···················· | ····················· | ···················· | ···················· | ···················· | |
| G | 8419 | hCoV-19/Brazil/AP-IEC-165513/2020\|EPI_ISL_524784\|2 | ···················· | ···················· | ····················· | ···················· | ···················· | ···················· | |
| G | 8420 | hCoV-19/Brazil/PA-IEC-165313/2020\|EPI_ISL_524785\|2 | ···················· | ···················· | ····················· | ···················· | ···················· | ···················· | |
| G | 8421 | hCoV-19/Brazil/PA-IEC-165302/2020\|EPI_ISL_524786\|2 | ···················· | ···················· | ····················· | ···················· | ···················· | ···················· | |
| G | 8422 | hCoV-19/Brazil/PA-IEC-163469/2020\|EPI_ISL_524787\|2 | ···················· | ···················· | ····················· | ···················· | ···················· | ···················· | |
| G | 8423 | hCoV-19/Brazil/MA-IEC-164827/2020\|EPI_ISL_524788\|2 | ···················· | ···················· | ····················· | ···················· | ···················· | ···················· | |
| G | 8424 | hCoV-19/Brazil/MA-IEC-166716/2020\|EPI_ISL_524789\|2 | ···················· | ···················· | ····················· | ···················· | ···················· | ···················· | |
| G | 8425 | hCoV-19/Brazil/MA-IEC-165398/2020\|EPI_ISL_524790\|2 | ···················· | ···················· | ····················· | ···················· | ···················· | ···················· | |
| G | 8426 | hCoV-19/Brazil/MA-IEC-162466/2020\|EPI_ISL_524791\|2 | ···················· | ···················· | ····················· | ···················· | ···················· | ···················· | |
| G | 8427 | hCoV-19/Brazil/AP-IEC-162620/2020\|EPI_ISL_524792\|2 | ···················· | ···················· | ····················· | ···················· | ···················· | ···················· | |
| G | 8428 | hCoV-19/Brazil/AP-IEC-165669/2020\|EPI_ISL_524793\|2 | ···················· | ···················· | ····················· | ···················· | ···················· | ···················· | |
| G | 8429 | hCoV-19/Brazil/MA-IEC-165425/2020\|EPI_ISL_524794\|2 | ···················· | ···················· | ····················· | ···················· | ···················· | ···················· | |
| G | 8430 | hCoV-19/Brazil/AP-IEC-163359/2020\|EPI_ISL_524795\|2 | ···················· | ···················· | ····················· | ···················· | ···················· | ···················· | |
| G | 8431 | hCoV-19/Brazil/AP-IEC-164920/2020\|EPI_ISL_524796\|2 | ···················· | ···················· | ····················· | ···················· | ···················· | ···················· | |
| G | 8432 | hCoV-19/Brazil/MA-IEC-162157/2020\|EPI_ISL_524797\|2 | ···················· | ···················· | ····················· | ···················· | ···················· | ···················· | |
| G | 8433 | hCoV-19/Brazil/RN-IEC-162277/2020\|EPI_ISL_524798\|2 | ···················· | ···················· | ····················· | ···················· | ···················· | ···················· | |
| G | 8434 | hCoV-19/Brazil/MA-IEC-166867/2020\|EPI_ISL_524799\|2 | ···················· | ···················· | ····················· | ···················· | ···················· | ···················· | |
| G | 8435 | hCoV-19/Switzerland/BS-UHB-42201335/2020\|EPI_ISL_5 | ···················· | ···················· | ····················· | ···················· | ···················· | ···················· | |
| G | 8436 | hCoV-19/Switzerland/BS-UHB-42201336/2020\|EPI_ISL_5 | ···················· | ···················· | ····················· | ···················· | ···················· | ···················· | |
| G | 8437 | hCoV-19/Switzerland/BS-UHB-42201408/2020\|EPI_ISL_5 | ···················· | ···················· | ····················· | ···················· | ···················· | ···················· | |
| G | 8438 | hCoV-19/Switzerland/BS-UHB-42201473/2020\|EPI_ISL_5 | ···················· | ···················· | ··················A·· | ···················· | ···················· | ···················· | 1 |
| G | 8439 | hCoV-19/Switzerland/BS-UHB-42201474/2020\|EPI_ISL_5 | ···················· | ···················· | ····················· | ···················· | ···················· | ···················· | |
| G | 8440 | hCoV-19/Switzerland/BS-UHB-42201508/2020\|EPI_ISL_5 | ···················· | ···················· | ····················· | ···················· | ···················· | ···················· | |
| G | 8441 | hCoV-19/Switzerland/BS-UHB-42201528/2020\|EPI_ISL_5 | ···················· | ···················· | ····················· | ···················· | ···················· | ···················· | |

| | | | | | | | | | | |
|---|---|---|---|---|---|---|---|---|---|---|
| G | 8442 | hCoV-19/Switzerland/BS-UHB-42202148/2020\|EPI_ISL_5 | ···················· | ···················· | ····················· | ···················· | ···················· | ···················· | | |
| G | 8443 | hCoV-19/Switzerland/BS-UHB-42202167/2020\|EPI_ISL_5 | ···················· | ···················· | ····················· | ···················· | ···················· | ···················· | | |
| G | 8444 | hCoV-19/England/BIRM-6384F/2020\|EPI_ISL_529650\|202 | ···················· | ···················· | ····················· | ···················· | ···················· | ···················· | | |
| G | 8445 | hCoV-19/England/BIRM-638A9/2020\|EPI_ISL_529651\|202 | ···················· | ···················· | ····················· | ···················· | ···················· | ···················· | | |
| G | 8446 | hCoV-19/England/BIRM-638C7/2020\|EPI_ISL_529652\|202 | ···················· | ···················· | ····················· | ···················· | ···················· | ···················· | | |
| G | 8447 | hCoV-19/England/BIRM-638F4/2020\|EPI_ISL_529653\|202 | ···················· | ···················· | ····················· | ···················· | ···················· | ···················· | | |
| G | 8448 | hCoV-19/England/BRIS-133C3C/2020\|EPI_ISL_529654\|20 | ···················· | ···················· | ····················· | ···················· | ···················· | ···················· | | |
| G | 8449 | hCoV-19/Wales/PHWC-169BB8/2020\|EPI_ISL_517600\|2020 | ···················· | ···················· | ····················· | ···················· | ···················· | ···················· | | |
| G | 8450 | hCoV-19/Wales/PHWC-169BD6/2020\|EPI_ISL_517601\|2020 | ···················· | ···················· | ····················· | ···················· | ···················· | ···················· | | |
| G | 8451 | hCoV-19/Wales/PHWC-169C1F/2020\|EPI_ISL_517605\|2020 | ···················· | ···················· | ····················· | ···················· | ·NNN················ | ···················· | | 1 |
| G | 8452 | hCoV-19/Wales/PHWC-169C3D/2020\|EPI_ISL_517607\|2020 | ···················· | ···················· | ····················· | ···················· | NNN················· | ···················· | | 1 |
| G | 8453 | hCoV-19/Wales/PHWC-169C5B/2020\|EPI_ISL_517609\|2020 | ···················· | ···················· | ····················· | ···················· | ···················· | ···················· | | |
| G | 8454 | hCoV-19/Germany/NI-UMCG-PE-S4/2020\|EPI_ISL_517611\| | ···················· | ···················· | ····················· | ···················· | ···················· | ···················· | | |
| G | 8455 | hCoV-19/Suriname/SR-02/2020\|EPI_ISL_517613\|2020-05 | ···················· | ···················· | ····················· | ···················· | ···················· | ···················· | | |
| G | 8456 | hCoV-19/Suriname/SR-04/2020\|EPI_ISL_517615\|2020-05 | ···················· | ···················· | ····················· | ···················· | ···················· | ···················· | | |
| G | 8457 | hCoV-19/Suriname/SR-05/2020\|EPI_ISL_517616\|2020-05 | ···················· | ···················· | ····················· | ···················· | ···················· | ···················· | | |
| G | 8458 | hCoV-19/Suriname/SR-09/2020\|EPI_ISL_517620\|2020-06 | ···················· | ···················· | ····················· | ···················· | ···················· | ···················· | | |
| G | 8459 | hCoV-19/Suriname/SR-10/2020\|EPI_ISL_517621\|2020-06 | ···················· | ···················· | ····················· | ···················· | ···················· | ···················· | | |
| G | 8460 | hCoV-19/Suriname/SR-11/2020\|EPI_ISL_517622\|2020-06 | ···················· | ···················· | ····················· | ···················· | ···················· | ···················· | | |
| G | 8461 | hCoV-19/Suriname/SR-12/2020\|EPI_ISL_517623\|2020-06 | ···················· | ···················· | ····················· | ···················· | ···················· | ···················· | | |
| G | 8462 | hCoV-19/Suriname/SR-13/2020\|EPI_ISL_517624\|2020-06 | ···················· | ···················· | ····················· | ···················· | ···················· | ···················· | | |
| G | 8463 | hCoV-19/Suriname/SR-14/2020\|EPI_ISL_517625\|2020-06 | ···············A···· | ···················· | ····················· | ···················· | ···················· | ···················· | | |
| G | 8464 | hCoV-19/Suriname/SR-15/2020\|EPI_ISL_517626\|2020-06 | ···················· | ···················· | ····················· | ···················· | ···················· | ···················· | | |
| G | 8465 | hCoV-19/Suriname/SR-16/2020\|EPI_ISL_517627\|2020-06 | ···················· | ···················· | ····················· | ···················· | ···················· | ···················· | | |
| G | 8466 | hCoV-19/Suriname/SR-17/2020\|EPI_ISL_517628\|2020-06 | ···················· | ···················· | ····················· | ···················· | ···················· | ···················· | | |
| G | 8467 | hCoV-19/Suriname/SR-18/2020\|EPI_ISL_517629\|2020-06 | ···················· | ···················· | ····················· | ···················· | ···················· | ···················· | | |
| G | 8468 | hCoV-19/Suriname/SR-19/2020\|EPI_ISL_517630\|2020-06 | ···················· | ···················· | ····················· | ···················· | ···················· | ···················· | | |
| G | 8469 | hCoV-19/Suriname/SR-20/2020\|EPI_ISL_517631\|2020-06 | ···················· | ···················· | ····················· | ···················· | ···················· | ···················· | | |
| G | 8470 | hCoV-19/Suriname/SR-22/2020\|EPI_ISL_517633\|2020-06 | ···················· | ···················· | ····················· | ···················· | ···················· | ···················· | | |
| G | 8471 | hCoV-19/Suriname/SR-25/2020\|EPI_ISL_517636\|2020-06 | ···················· | ···················· | ····················· | ···················· | ···················· | ···················· | | |
| G | 8472 | hCoV-19/Suriname/SR-26/2020\|EPI_ISL_517637\|2020-06 | ···················· | ···················· | ····················· | ···················· | ···················· | ···················· | | |
| G | 8473 | hCoV-19/Suriname/SR-27/2020\|EPI_ISL_517638\|2020-06 | ···················· | ···················· | ····················· | ···················· | ···················· | ···················· | | |
| G | 8474 | hCoV-19/Suriname/SR-28/2020\|EPI_ISL_517639\|2020-06 | ···················· | ···················· | ····················· | ···················· | ···················· | ···················· | | |
| G | 8475 | hCoV-19/Suriname/SR-29/2020\|EPI_ISL_517640\|2020-06 | ···················· | ···················· | ····················· | ···················· | ···················· | ···················· | | |
| G | 8476 | hCoV-19/Suriname/SR-30/2020\|EPI_ISL_517641\|2020-06 | ···················· | ···················· | ····················· | ···················· | ···················· | ···················· | | |
| G | 8477 | hCoV-19/Suriname/SR-33/2020\|EPI_ISL_517644\|2020-06 | ···················· | ···················· | ····················· | ···················· | ···················· | ···················· | | |
| G | 8478 | hCoV-19/Suriname/SR-34/2020\|EPI_ISL_517645\|2020-06 | ···················· | ···················· | ····················· | ···················· | ···················· | ···················· | | |
| G | 8479 | hCoV-19/Suriname/SR-35/2020\|EPI_ISL_517646\|2020-06 | ···················· | ···················· | ····················· | ···················· | ···················· | ···················· | | |
| G | 8480 | hCoV-19/Suriname/SR-36/2020\|EPI_ISL_517647\|2020-06 | ···················· | ···················· | ····················· | ···················· | ···················· | ···················· | | |
| G | 8481 | hCoV-19/Suriname/SR-37/2020\|EPI_ISL_517648\|2020-06 | ···················· | ···················· | ····················· | ···················· | ···················· | ···················· | | |
| G | 8482 | hCoV-19/Suriname/SR-39/2020\|EPI_ISL_517650\|2020-06 | ···················· | ···················· | ····················· | ···················· | ···················· | ···················· | | |
| G | 8483 | hCoV-19/Suriname/SR-40/2020\|EPI_ISL_517651\|2020-06 | ···················· | ···················· | ····················· | ···················· | ···················· | ···················· | | |
| G | 8484 | hCoV-19/Suriname/SR-41/2020\|EPI_ISL_517652\|2020-06 | ···················· | ···················· | ····················· | ···················· | ···················· | ···················· | | |
| G | 8485 | hCoV-19/Suriname/SR-42/2020\|EPI_ISL_517653\|2020-06 | ···················· | ···················· | ····················· | ···················· | ···················· | ···················· | | |
| G | 8486 | hCoV-19/Suriname/SR-43/2020\|EPI_ISL_517654\|2020-06 | ···················· | ···················· | ····················· | ···················· | ···················· | ···················· | | |
| G | 8487 | hCoV-19/Suriname/SR-46/2020\|EPI_ISL_517657\|2020-07 | ···················· | ···················· | ····················· | ···················· | ···················· | ···················· | | |
| G | 8488 | hCoV-19/Suriname/SR-47/2020\|EPI_ISL_517658\|2020-07 | ···················· | ···················· | ····················· | ···················· | ···················· | ···················· | | |
| G | 8489 | hCoV-19/Suriname/SR-48/2020\|EPI_ISL_517659\|2020-07 | ···················· | ···················· | ····················· | ···················· | ···················· | ···················· | | |
| G | 8490 | hCoV-19/Australia/VIC4516/2020\|EPI_ISL_518927\|2020 | ···················· | ···················· | ····················· | ···················· | ···················· | ···················· | | |
| G | 8491 | hCoV-19/Australia/VIC4517/2020\|EPI_ISL_518928\|2020 | ···················· | ···················· | ····················· | ···················· | ···················· | ···················· | | |
| G | 8492 | hCoV-19/Australia/VIC4518/2020\|EPI_ISL_518929\|2020 | ··················T· | ···················· | ····················· | ···················· | ···················· | ···················· | 1 | |
| G | 8493 | hCoV-19/Australia/VIC4519/2020\|EPI_ISL_518930\|2020 | ···················· | ···················· | ····················· | ···················· | ···················· | ···················· | | |
| G | 8494 | hCoV-19/Australia/VIC4520/2020\|EPI_ISL_518931\|2020 | ···················· | ···················· | ····················· | ···················· | ···················· | ···················· | | |
| G | 8495 | hCoV-19/Australia/VIC4521/2020\|EPI_ISL_518932\|2020 | ···················· | ···················· | ····················· | ···················· | ···················· | ···················· | | |
| G | 8496 | hCoV-19/Australia/VIC4522/2020\|EPI_ISL_518933\|2020 | ···················· | ···················· | ····················· | ···················· | ···················· | ···················· | | |
| G | 8497 | hCoV-19/Australia/VIC4523/2020\|EPI_ISL_518934\|2020 | ···················· | ···················· | ····················· | ···················· | ···················· | ···················· | | |
| G | 8498 | hCoV-19/Australia/VIC4526/2020\|EPI_ISL_518937\|2020 | ···················· | ···················· | ····················· | ···················· | ···················· | ···················· | | |
| G | 8499 | hCoV-19/Australia/VIC4527/2020\|EPI_ISL_518938\|2020 | ···················· | ···················· | ····················· | ···················· | ···················· | ···················· | | |
| G | 8500 | hCoV-19/Australia/VIC4528/2020\|EPI_ISL_518939\|2020 | ···················· | ···················· | ····················· | ···················· | ···················· | ···················· | | |
| G | 8501 | hCoV-19/Australia/VIC4529/2020\|EPI_ISL_518940\|2020 | ···················· | ···················· | ····················· | ···················· | ···················· | ···················· | | |
| G | 8502 | hCoV-19/Australia/VIC4530/2020\|EPI_ISL_518941\|2020 | ···················· | ···················· | ····················· | ···················· | ···················· | ···················· | | |
| G | 8503 | hCoV-19/Australia/VIC4532/2020\|EPI_ISL_518943\|2020 | ···················· | ···················· | ····················· | ···················· | ···················· | ···················· | | |
| G | 8504 | hCoV-19/Australia/VIC4533/2020\|EPI_ISL_518944\|2020 | ···················· | ···················· | ····················· | ···················· | ···················· | ···················· | | |
| G | 8505 | hCoV-19/Australia/VIC4534/2020\|EPI_ISL_518945\|2020 | ···················· | ···················· | ····················· | ···················· | ···················· | ···················· | | |
| G | 8506 | hCoV-19/Australia/VIC4535/2020\|EPI_ISL_518946\|2020 | ···················· | ···················· | ····················· | ···················· | ···················· | ···················· | | |
| G | 8507 | hCoV-19/Australia/VIC4536/2020\|EPI_ISL_518947\|2020 | ···················· | ···················· | ····················· | ···················· | ···················· | ···················· | | |
| G | 8508 | hCoV-19/Australia/VIC4537/2020\|EPI_ISL_518948\|2020 | ···················· | ···················· | ····················· | ···················· | ···················· | ···················· | | |
| G | 8509 | hCoV-19/Australia/VIC4538/2020\|EPI_ISL_518949\|2020 | ···················· | ···················· | ····················· | ···················· | ···················· | ···················· | | |
| G | 8510 | hCoV-19/Australia/VIC4539/2020\|EPI_ISL_518950\|2020 | ···················· | ···················· | ····················· | ···················· | ···················· | ···················· | | |
| G | 8511 | hCoV-19/Australia/VIC4540/2020\|EPI_ISL_518951\|2020 | ···················· | ···················· | ····················· | ···················· | ···················· | ···················· | | |
| G | 8512 | hCoV-19/Australia/VIC4541/2020\|EPI_ISL_518952\|2020 | ···················· | ···················· | ····················· | ···················· | ···················· | ···················· | | |
| G | 8513 | hCoV-19/Australia/VIC4543/2020\|EPI_ISL_518954\|2020 | ···················· | ···················· | ····················· | ···················· | ···················· | ···················· | | |
| G | 8514 | hCoV-19/Australia/VIC4544/2020\|EPI_ISL_518955\|2020 | ···················· | ···················· | ····················· | ···················· | ···················· | ···················· | | |
| G | 8515 | hCoV-19/Australia/VIC4545/2020\|EPI_ISL_518956\|2020 | ···················· | ···················· | ····················· | ···················· | ···················· | ···················· | | |
| G | 8516 | hCoV-19/Australia/VIC4546/2020\|EPI_ISL_518957\|2020 | ···················· | ···················· | ····················· | ···················· | ···················· | ···················· | | |
| G | 8517 | hCoV-19/Australia/VIC4547/2020\|EPI_ISL_518958\|2020 | ···················· | ···················· | ····················· | ···················· | ···················· | ···················· | | |
| G | 8518 | hCoV-19/Australia/VIC4548/2020\|EPI_ISL_518959\|2020 | ···················· | ···················· | ····················· | ···················· | ···················· | ···················· | | |
| G | 8519 | hCoV-19/Australia/VIC4549/2020\|EPI_ISL_518960\|2020 | ···················· | ···················· | ····················· | ···················· | ···················· | ···················· | | |
| G | 8520 | hCoV-19/Australia/VIC4550/2020\|EPI_ISL_518961\|2020 | ···················· | ···················· | ····················· | ···················· | ···················· | ···················· | | |
| G | 8521 | hCoV-19/Australia/VIC4552/2020\|EPI_ISL_518962\|2020 | ···················· | ···················· | ····················· | ···················· | ···················· | ···················· | | |

| | | | | | | | | |
|---|---|---|---|---|---|---|---|---|
| G | 8522 | hCoV-19/Australia/VIC4553/2020\|EPI_ISL_518963\|2020 | ···················· | ···················· | ···················· | ···················· | ···················· | ···················· |
| G | 8523 | hCoV-19/Australia/VIC4554/2020\|EPI_ISL_518964\|2020 | ···················· | ···················· | ···················· | ···················· | ···················· | ···················· |
| G | 8524 | hCoV-19/Australia/VIC4555/2020\|EPI_ISL_518965\|2020 | ···················· | ···················· | ···················· | ···················· | ···················· | ···················· |
| G | 8525 | hCoV-19/Australia/VIC4556/2020\|EPI_ISL_518966\|2020 | ···················· | ···················· | ···················· | ···················· | ···················· | ···················· |
| G | 8526 | hCoV-19/Australia/VIC4557/2020\|EPI_ISL_518967\|2020 | ···················· | ···················· | ···················· | ···················· | ···················· | ···················· |
| G | 8527 | hCoV-19/Australia/VIC4558/2020\|EPI_ISL_518968\|2020 | ···················· | ···················· | ···················· | ···················· | ···················· | ···················· |
| G | 8528 | hCoV-19/Australia/VIC4559/2020\|EPI_ISL_518969\|2020 | ···················· | ···················· | ···················· | ···················· | ···················· | ···················· |
| G | 8529 | hCoV-19/Australia/VIC4561/2020\|EPI_ISL_518971\|2020 | ···················· | ···················· | ···················· | ···················· | ···················· | ···················· |
| G | 8530 | hCoV-19/Australia/VIC4562/2020\|EPI_ISL_518972\|2020 | ···················· | ···················· | ···················· | ···················· | ···················· | ···················· |
| G | 8531 | hCoV-19/Australia/VIC4563/2020\|EPI_ISL_518973\|2020 | ···················· | ···················· | ···················· | ···················· | ···················· | ···················· |
| G | 8532 | hCoV-19/Australia/VIC4565/2020\|EPI_ISL_518975\|2020 | ···················· | ···················· | ···················· | ···················· | ···················· | ···················· |
| G | 8533 | hCoV-19/Australia/VIC4567/2020\|EPI_ISL_518976\|2020 | ···················· | ···················· | ···················· | ···················· | ···················· | ···················· |
| G | 8534 | hCoV-19/Australia/VIC4571/2020\|EPI_ISL_518977\|2020 | ···················· | ···················· | ···················· | ···················· | ···················· | ···················· |
| G | 8535 | hCoV-19/Australia/VIC4572/2020\|EPI_ISL_518978\|2020 | ···················· | ···················· | ···················· | ···················· | ···················· | ···················· |
| G | 8536 | hCoV-19/Australia/VIC4574/2020\|EPI_ISL_518979\|2020 | ···················· | ···················· | ···················· | ···················· | ···················· | ···················· |
| G | 8537 | hCoV-19/Australia/VIC4576/2020\|EPI_ISL_518980\|2020 | ···················· | ···················· | ···················· | ···················· | ···················· | ···················· |
| G | 8538 | hCoV-19/Australia/VIC4579/2020\|EPI_ISL_518981\|2020 | ···················· | ···················· | ···················· | ···················· | ···················· | ···················· |
| G | 8539 | hCoV-19/Australia/VIC4580/2020\|EPI_ISL_518982\|2020 | ···················· | ···················· | ···················· | ···················· | ···················· | ···················· |
| G | 8540 | hCoV-19/Australia/VIC4581/2020\|EPI_ISL_518983\|2020 | ···················· | ···················· | ···················· | ···················· | ···················· | ···················· |
| G | 8541 | hCoV-19/Australia/VIC4583/2020\|EPI_ISL_518984\|2020 | ···················· | ···················· | ···················· | ···················· | ···················· | ···················· |
| G | 8542 | hCoV-19/Australia/VIC4584/2020\|EPI_ISL_518985\|2020 | ···················· | ···················· | ···················· | ···················· | ···················· | ···················· |
| G | 8543 | hCoV-19/Australia/VIC4585/2020\|EPI_ISL_518986\|2020 | ···················· | ···················· | ···················· | ···················· | ···················· | ···················· |
| G | 8544 | hCoV-19/Australia/VIC4586/2020\|EPI_ISL_518987\|2020 | ···················· | ···················· | ···················· | ···················· | ···················· | ···················· |
| G | 8545 | hCoV-19/Australia/VIC4587/2020\|EPI_ISL_518988\|2020 | ···················· | ···················· | ···················· | ···················· | ···················· | ···················· |
| G | 8546 | hCoV-19/Australia/VIC4588/2020\|EPI_ISL_518989\|2020 | ···················· | ···················· | ···················· | ···················· | ···················· | ···················· |
| G | 8547 | hCoV-19/Australia/VIC4589/2020\|EPI_ISL_518990\|2020 | ···················· | ···················· | ···················· | ···················· | ···················· | ···················· |
| G | 8548 | hCoV-19/Australia/VIC4590/2020\|EPI_ISL_518991\|2020 | ···················· | ···················· | ···················· | ···················· | ···················· | ···················· |
| G | 8549 | hCoV-19/Australia/VIC4591/2020\|EPI_ISL_518992\|2020 | ···················· | ···················· | ···················· | ···················· | ···················· | ···················· |
| G | 8550 | hCoV-19/Australia/VIC4592/2020\|EPI_ISL_518993\|2020 | ···················· | ···················· | ···················· | ···················· | ···················· | ···················· |
| G | 8551 | hCoV-19/Australia/VIC4593/2020\|EPI_ISL_518994\|2020 | ···················· | ···················· | ···················· | ···················· | ···················· | ···················· |
| G | 8552 | hCoV-19/Australia/VIC4594/2020\|EPI_ISL_518995\|2020 | ···················· | ···················· | ···················· | ···················· | ···················· | ···················· |
| G | 8553 | hCoV-19/Australia/VIC4595/2020\|EPI_ISL_518996\|2020 | ···················· | ···················· | ···················· | ···················· | ···················· | ···················· |
| G | 8554 | hCoV-19/Australia/VIC4596/2020\|EPI_ISL_518997\|2020 | ···················· | ···················· | ···················· | ···················· | ···················· | ···················· |
| G | 8555 | hCoV-19/Australia/VIC4597/2020\|EPI_ISL_518998\|2020 | ···················· | ···················· | ···················· | ···················· | ···················· | ···················· |
| G | 8556 | hCoV-19/Australia/VIC4598/2020\|EPI_ISL_518999\|2020 | ···················· | ···················· | ···················· | ···················· | ···················· | ···················· |
| G | 8557 | hCoV-19/Australia/VIC2494/2020\|EPI_ISL_522192\|2020 | ···················· | ···················· | ···················· | ···················· | ···················· | ···················· |
| G | 8558 | hCoV-19/Australia/VIC2495/2020\|EPI_ISL_522193\|2020 | ···················· | ···················· | ···················· | ···················· | ···················· | ···················· |
| G | 8559 | hCoV-19/Australia/VIC2496/2020\|EPI_ISL_522194\|2020 | ···················· | ···················· | ···················· | ···················· | ···················· | ···················· |
| G | 8560 | hCoV-19/Australia/VIC2497/2020\|EPI_ISL_522195\|2020 | ···················· | ···················· | ···················· | ···················· | ···················· | ···················· |
| G | 8561 | hCoV-19/Australia/VIC2499/2020\|EPI_ISL_522196\|2020 | ···················· | ···················· | ···················· | ···················· | ···················· | ···················· |
| G | 8562 | hCoV-19/Australia/VIC2500/2020\|EPI_ISL_522197\|2020 | ···················· | ···················· | ···················· | ···················· | ···················· | ···················· |
| G | 8563 | hCoV-19/Australia/VIC2501/2020\|EPI_ISL_522198\|2020 | ···················· | ···················· | ···················· | ···················· | ···················· | ···················· |
| G | 8564 | hCoV-19/Australia/VIC2502/2020\|EPI_ISL_522199\|2020 | ···················· | ···················· | ···················· | ···················· | ···················· | ···················· |
| G | 8565 | hCoV-19/Brazil/PA-IEC-166687/2020\|EPI_ISL_524800\|2 | ···················· | ···················· | ···················· | ···················· | ···················· | ···················· |
| G | 8566 | hCoV-19/Australia/NSW2510/2020\|EPI_ISL_527030\|2020 | ···················· | ···················· | ···················· | ···················· | ···················· | ···················· |
| G | 8567 | hCoV-19/Australia/NSW2511/2020\|EPI_ISL_527031\|2020 | ···················· | ···················· | ···················· | ···················· | ···················· | ···················· |
| G | 8568 | hCoV-19/Australia/NSW2512/2020\|EPI_ISL_527032\|2020 | ···················· | ···················· | ···················· | ···················· | ···················· | ···················· |
| G | 8569 | hCoV-19/Australia/NSW2513/2020\|EPI_ISL_527033\|2020 | ···················· | ·A·················· | ···················· | ···················· | ···················· | ··A················· |
| G | 8570 | hCoV-19/Australia/NSW2515/2020\|EPI_ISL_527034\|2020 | ···················· | ·A·················· | ···················· | ···················· | ···················· | ··A················· |
| G | 8571 | hCoV-19/Australia/NSW2519/2020\|EPI_ISL_527035\|2020 | ···················· | ···················· | ···················· | ···················· | ···················· | ···················· |
| G | 8572 | hCoV-19/Australia/NSW2526/2020\|EPI_ISL_527036\|2020 | ···················· | ···················· | ···················· | ···················· | ···················· | ···················· |
| G | 8573 | hCoV-19/Australia/NSW2533/2020\|EPI_ISL_527037\|2020 | ···················· | ···················· | ···················· | ···················· | ···················· | ···················· |
| G | 8574 | hCoV-19/Australia/NSW2534/2020\|EPI_ISL_527038\|2020 | ···················· | ···················· | ···················· | ···················· | ···················· | ···················· |
| G | 8575 | hCoV-19/Australia/NSW2538/2020\|EPI_ISL_527040\|2020 | ···················· | ···················· | ···················· | ···················· | ···················· | ···················· |
| G | 8576 | hCoV-19/Australia/NSW2539/2020\|EPI_ISL_527041\|2020 | ···················· | ···················· | ···················· | ···················· | ···················· | ···················· |
| G | 8577 | hCoV-19/Australia/NSW2541/2020\|EPI_ISL_527042\|2020 | ···················· | ···················· | ···················· | ···················· | ···················· | ···················· |
| G | 8578 | hCoV-19/Australia/NSW2542/2020\|EPI_ISL_527043\|2020 | ···················· | ···················· | ···················· | ···················· | ···················· | ···················· |
| G | 8579 | hCoV-19/Australia/NSW2543/2020\|EPI_ISL_527044\|2020 | ···················· | ···················· | ···················· | ···················· | ···················· | ···················· |
| G | 8580 | hCoV-19/Australia/NSW2544/2020\|EPI_ISL_527045\|2020 | ···················· | ···················· | ···················· | ···················· | ···················· | ···················· |
| G | 8581 | hCoV-19/Australia/NSW2545/2020\|EPI_ISL_527046\|2020 | ···················· | ···················· | ···················· | ···················· | ···················· | ···················· |
| G | 8582 | hCoV-19/Australia/NSW2546/2020\|EPI_ISL_527047\|2020 | ···················· | ···················· | ···················· | ···················· | ···················· | ···················· |
| G | 8583 | hCoV-19/Australia/NSW2547/2020\|EPI_ISL_527048\|2020 | ···················· | ·A·················· | ···················· | ···················· | ···················· | ···················· |
| G | 8584 | hCoV-19/Australia/NSW2548/2020\|EPI_ISL_527049\|2020 | ···················· | ···················· | ···················· | ···················· | ···················· | ···················· |
| G | 8585 | hCoV-19/Australia/NSW2549/2020\|EPI_ISL_527050\|2020 | ···················· | ···················· | ···················· | ···················· | ···················· | ···················· |
| G | 8586 | hCoV-19/Australia/NSW2550/2020\|EPI_ISL_527051\|2020 | ···················· | ···················· | ···················· | ···················· | ···················· | ···················· |
| G | 8587 | hCoV-19/Australia/NSW2551/2020\|EPI_ISL_527052\|2020 | ···················· | ···················· | ···················· | ···················· | ···················· | ···················· |
| G | 8588 | hCoV-19/Australia/NSW2552/2020\|EPI_ISL_527053\|2020 | ···················· | ···················· | ···················· | ···················· | ···················· | ···················· |
| G | 8589 | hCoV-19/Australia/NSW2557/2020\|EPI_ISL_527054\|2020 | ···················· | ···················· | ···················· | ···················· | ···················· | ···················· |
| G | 8590 | hCoV-19/Australia/NSW2562/2020\|EPI_ISL_527055\|2020 | ···················· | ···················· | ···················· | ···················· | ···················· | ···················· |
| G | 8591 | hCoV-19/Australia/NSW2563/2020\|EPI_ISL_527056\|2020 | ···················· | ···················· | ···················· | ···················· | ···················· | ···················· |
| G | 8592 | hCoV-19/Australia/NSW2570/2020\|EPI_ISL_527057\|2020 | ···················· | ···················· | ···················· | ···················· | ···················· | ···················· |
| G | 8593 | hCoV-19/Australia/NSW2571/2020\|EPI_ISL_527058\|2020 | ···················· | ···················· | ···················· | ···················· | ···················· | ···················· |
| G | 8594 | hCoV-19/Australia/NSW2573/2020\|EPI_ISL_527059\|2020 | ···················· | ···················· | ···················· | ···················· | ···················· | ···················· |
| G | 8595 | hCoV-19/Australia/NSW2574/2020\|EPI_ISL_527060\|2020 | ···················· | ···················· | ···················· | ···················· | ···················· | ···················· |
| G | 8596 | hCoV-19/Australia/NSW2576/2020\|EPI_ISL_527061\|2020 | ···················· | ···················· | ···················· | ···················· | ···················· | ···················· |
| G | 8597 | hCoV-19/Australia/NSW2577/2020\|EPI_ISL_527062\|2020 | ···················· | ·A·················· | ···················· | ···················· | ···················· | ···················· |
| G | 8598 | hCoV-19/Australia/NSW2616/2020\|EPI_ISL_527063\|2020 | ···················· | ···················· | ···················· | ···················· | ···················· | ···················· |
| G | 8599 | hCoV-19/Australia/NSW2647/2020\|EPI_ISL_527064\|2020 | ···················· | ···················· | ···················· | ···················· | ···················· | ···················· |
| G | 8600 | hCoV-19/Switzerland/BL-UHB-42201578/2020\|EPI_ISL_5 | ···················· | ···················· | ···················· | ···················· | ·NN················· | ···················· |
| G | 8601 | hCoV-19/Switzerland/BS-UHB-42201579/2020\|EPI_ISL_5 | ···················· | ···················· | ···················· | ···················· | ···················· | ···················· |

| | | | | | | | | |
|---|---|---|---|---|---|---|---|---|
| G | 8602 | hCoV-19/Switzerland/BS-UHB-42201625/2020\|EPI_ISL_5 | ···················· | ···················· | ···················· | ···················· | ···················· | ···················· |
| G | 8603 | hCoV-19/Switzerland/BS-UHB-42201738/2020\|EPI_ISL_5 | ···················· | ···················· | ···················· | ···················· | ···················· | ···················· |
| G | 8604 | hCoV-19/Switzerland/BS-UHB-42201769/2020\|EPI_ISL_5 | ···················· | ···················· | ···················· | ···················· | ···················· | ···················· |
| G | 8605 | hCoV-19/Switzerland/BS-UHB-42201771/2020\|EPI_ISL_5 | ···················· | ···················· | ···················· | ···················· | ···················· | ···················· |
| G | 8606 | hCoV-19/Switzerland/BL-UHB-42201781/2020\|EPI_ISL_5 | ···················· | ···················· | ···················· | ···················· | ···················· | ···················· |
| G | 8607 | hCoV-19/Switzerland/BS-UHB-42202026/2020\|EPI_ISL_5 | ···················· | ···················· | ···················· | ···················· | ···················· | ···················· |
| G | 8608 | hCoV-19/Switzerland/BS-UHB-42202033/2020\|EPI_ISL_5 | ···················· | ···················· | ···················· | ···················· | ···················· | ···················· |
| G | 8609 | hCoV-19/Switzerland/BS-UHB-42202078/2020\|EPI_ISL_5 | ···················· | ···················· | ···················· | ···················· | ···················· | ···················· |
| G | 8610 | hCoV-19/Switzerland/BS-UHB-42202140/2020\|EPI_ISL_5 | ···················· | ···················· | ···················· | ···················· | ···················· | ···················· |
| G | 8611 | hCoV-19/Switzerland/BS-UHB-42202147/2020\|EPI_ISL_5 | ···················· | ···················· | ···················· | ···················· | ···················· | ···················· |
| G | 8612 | hCoV-19/Switzerland/BS-UHB-42202218/2020\|EPI_ISL_5 | ···················· | ···················· | ···················· | ···················· | ···················· | ···················· |
| G | 8613 | hCoV-19/Switzerland/BS-UHB-42202238/2020\|EPI_ISL_5 | ···················· | ···················· | ···················· | ···················· | ·NN················· | ···················· |
| G | 8614 | hCoV-19/Switzerland/BS-UHB-42202239/2020\|EPI_ISL_5 | ···················· | ···················· | ···················· | ···················· | ···················· | ···················· |
| G | 8616 | hCoV-19/Switzerland/BS-UHB-42203612/2020\|EPI_ISL_5 | ···················· | ···················· | ···················· | ···················· | ···················· | ···················· |
| G | 8617 | hCoV-19/Switzerland/BS-UHB-42203645/2020\|EPI_ISL_5 | ···················· | ···················· | ···················· | ···················· | ···················· | ···················· |
| G | 8618 | hCoV-19/Switzerland/BS-UHB-42203656/2020\|EPI_ISL_5 | ···················· | ···················· | ···················· | ···················· | ···················· | ···················· |
| G | 8619 | hCoV-19/Switzerland/BS-UHB-42203665/2020\|EPI_ISL_5 | ···················· | ···················· | ···················· | ···················· | ···················· | ···················· |
| G | 8620 | hCoV-19/Switzerland/BS-UHB-42203696/2020\|EPI_ISL_5 | ···················· | ···················· | ···················· | ···················· | ···················· | ···················· |
| G | 8621 | hCoV-19/Switzerland/BS-UHB-42203703/2020\|EPI_ISL_5 | ···················· | ···················· | ···················· | ···················· | ···················· | ···················· |
| G | 8622 | hCoV-19/Switzerland/BS-UHB-42203738/2020\|EPI_ISL_5 | ···················· | ···················· | ···················· | ···················· | ···················· | ···················· |
| G | 8623 | hCoV-19/Switzerland/BL-UHB-42203765/2020\|EPI_ISL_5 | ···················· | ···················· | ···················· | ···················· | ···················· | ···················· |
| G | 8624 | hCoV-19/Switzerland/BS-UHB-42203813/2020\|EPI_ISL_5 | ···················· | ···················· | ···················· | ···················· | ···················· | ···················· |
| G | 8625 | hCoV-19/Switzerland/BS-UHB-42204170/2020\|EPI_ISL_5 | ···················· | ···················· | ···················· | ···················· | ···················· | ···················· |
| G | 8626 | hCoV-19/Switzerland/BS-UHB-42204989/2020\|EPI_ISL_5 | ···················· | ···················· | ···················· | ···················· | ···················· | ···················· |
| G | 8628 | hCoV-19/Switzerland/BS-UHB-65238285/2020\|EPI_ISL_5 | ···················· | ···················· | ···················· | ···················· | ···················· | ···················· |
| G | 8629 | hCoV-19/Switzerland/BS-UHB-65238783/2020\|EPI_ISL_5 | ···················· | ···················· | ···················· | ···················· | ···················· | ···················· |
| G | 8630 | hCoV-19/India/HR-THSTI2010A/2020\|EPI_ISL_528382\|20 | ···················· | ···················· | ···················· | ···················· | ···················· | ···················· |
| G | 8631 | hCoV-19/India/HR-THSTI2010B/2020\|EPI_ISL_528383\|20 | ···················· | ···················· | ···················· | ···················· | ···················· | ···················· |
| G | 8632 | hCoV-19/India/HR-THSTI2010C/2020\|EPI_ISL_528384\|20 | ···················· | ···················· | ···················· | ···················· | ···················· | ···················· |
| G | 8633 | hCoV-19/India/HR-THSTI2010D/2020\|EPI_ISL_528385\|20 | ···················· | ···················· | ···················· | ···················· | ···················· | ···················· |
| G | 8634 | hCoV-19/Egypt/C-VSVRI-SERVAC/2020\|EPI_ISL_528386\|2 | ···················· | ···················· | ···················· | ···················· | ···················· | ···················· |
| G | 8635 | hCoV-19/England/20168039704/2020\|EPI_ISL_528388\|20 | ···················· | ···················· | ···················· | ···················· | ···················· | ···················· |
| G | 8636 | hCoV-19/England/20168039904/2020\|EPI_ISL_528389\|20 | ···················· | ···················· | ···················· | ···················· | ···················· | ···················· |
| G | 8637 | hCoV-19/England/20189113304/2020\|EPI_ISL_528391\|20 | ···················· | ···················· | ···················· | ···················· | ···················· | ···················· |
| G | 8638 | hCoV-19/England/20204114504/2020\|EPI_ISL_528392\|20 | ···················· | ···················· | ···················· | ···················· | ···················· | ···················· |
| G | 8639 | hCoV-19/England/20166007904/2020\|EPI_ISL_528398\|20 | ···················· | ···················· | ···················· | ···················· | ···················· | ···················· |
| G | 8640 | hCoV-19/England/20166019004/2020\|EPI_ISL_528399\|20 | ···················· | ···················· | ···················· | ···················· | ···················· | ···················· |
| G | 8641 | hCoV-19/England/BIRM-61BA2/2020\|EPI_ISL_529600\|202 | ···················· | ···················· | ···················· | ···················· | ···················· | ···················· |
| G | 8642 | hCoV-19/England/BIRM-61BFD/2020\|EPI_ISL_529601\|202 | ···················· | ···················· | ···················· | ···················· | ···················· | ···················· |
| G | 8643 | hCoV-19/England/BIRM-61C27/2020\|EPI_ISL_529602\|202 | ···················· | ···················· | ···················· | ···················· | ···················· | ···················· |
| G | 8644 | hCoV-19/England/BIRM-61C45/2020\|EPI_ISL_529603\|202 | ···················· | ···················· | ···················· | ···················· | ···················· | ···················· |
| G | 8645 | hCoV-19/England/BIRM-61CFA/2020\|EPI_ISL_529604\|202 | ···················· | ···················· | ···················· | ···················· | ···················· | ···················· |
| G | 8646 | hCoV-19/England/BIRM-61D06/2020\|EPI_ISL_529605\|202 | ···················· | ···················· | ···················· | ···················· | ···················· | ···················· |
| G | 8647 | hCoV-19/England/BIRM-61D15/2020\|EPI_ISL_529606\|202 | ···················· | ···················· | ···················· | ···················· | ···················· | ···················· |
| G | 8648 | hCoV-19/England/BIRM-61D51/2020\|EPI_ISL_529607\|202 | ···················· | ···················· | ···················· | ···················· | ···················· | ···················· |
| G | 8649 | hCoV-19/England/BIRM-61D7F/2020\|EPI_ISL_529608\|202 | ···················· | ···················· | ···················· | ···················· | ···················· | ···················· |
| G | 8650 | hCoV-19/England/BIRM-61D8E/2020\|EPI_ISL_529609\|202 | ···················· | ···················· | ···················· | ···················· | ···················· | ···················· |
| G | 8651 | hCoV-19/England/BIRM-61DE8/2020\|EPI_ISL_529610\|202 | ···················· | ···················· | ···················· | ···················· | ···················· | ···················· |
| G | 8652 | hCoV-19/England/BIRM-61E12/2020\|EPI_ISL_529611\|202 | ···················· | ···················· | ···················· | ···················· | ···················· | ···················· |
| G | 8653 | hCoV-19/England/BIRM-61E21/2020\|EPI_ISL_529612\|202 | ···················· | ···················· | ···················· | ···················· | ···················· | ···················· |
| G | 8654 | hCoV-19/England/BIRM-61E8B/2020\|EPI_ISL_529613\|202 | ···················· | ···················· | ···················· | ···················· | ···················· | ···················· |
| G | 8655 | hCoV-19/England/BIRM-61E9A/2020\|EPI_ISL_529614\|202 | ···················· | ···················· | ···················· | ···················· | ···················· | ···················· |
| G | 8656 | hCoV-19/England/BIRM-61EB8/2020\|EPI_ISL_529615\|202 | ···················· | ···················· | ···················· | ···················· | ···················· | ···················· |
| G | 8657 | hCoV-19/England/BIRM-61ED6/2020\|EPI_ISL_529616\|202 | ···················· | ···················· | ···················· | ···················· | ···················· | ···················· |
| G | 8658 | hCoV-19/England/BIRM-61F1F/2020\|EPI_ISL_529617\|202 | ···················· | ···················· | ···················· | ···················· | ···················· | ···················· |
| G | 8659 | hCoV-19/England/BIRM-62682/2020\|EPI_ISL_529618\|202 | ···················· | ···················· | ···················· | ···················· | ···················· | ···················· |
| G | 8660 | hCoV-19/England/BIRM-62691/2020\|EPI_ISL_529619\|202 | ···················· | ···················· | ···················· | ···················· | ···················· | ···················· |
| G | 8661 | hCoV-19/England/BIRM-626CE/2020\|EPI_ISL_529620\|202 | ···················· | ···················· | ···················· | ···················· | ···················· | ···················· |
| G | 8662 | hCoV-19/England/BIRM-62707/2020\|EPI_ISL_529621\|202 | ···················· | ···················· | ···················· | ···················· | ···················· | ···················· |
| G | 8663 | hCoV-19/England/BIRM-62813/2020\|EPI_ISL_529622\|202 | ···················· | ···················· | ···················· | ···················· | ···················· | ···················· |
| G | 8664 | hCoV-19/England/BIRM-62831/2020\|EPI_ISL_529623\|202 | ···················· | ·A·················· | ···················· | ···················· | ···················· | ···A················ |
| G | 8665 | hCoV-19/England/BIRM-6285F/2020\|EPI_ISL_529624\|202 | ···················· | ···················· | ···················· | ···················· | ···················· | ···················· |
| G | 8666 | hCoV-19/England/BIRM-6287D/2020\|EPI_ISL_529625\|202 | ···················· | ···················· | ···················· | ···················· | ···················· | ···················· |
| G | 8667 | hCoV-19/England/BIRM-628C8/2020\|EPI_ISL_529626\|202 | ···················· | ·A·················· | ···················· | ···················· | ···················· | ···A················ |
| G | 8668 | hCoV-19/England/BIRM-628E6/2020\|EPI_ISL_529627\|202 | ···················· | ···················· | ···················· | ···················· | ···················· | ···················· |
| G | 8669 | hCoV-19/England/BIRM-62901/2020\|EPI_ISL_529628\|202 | ···················· | ·A·················· | ···················· | ···················· | ···················· | ···················· |
| G | 8670 | hCoV-19/England/BIRM-6292F/2020\|EPI_ISL_529629\|202 | ···················· | ·A·················· | ···················· | ···················· | ···················· | ···A················ |
| G | 8671 | hCoV-19/England/BIRM-6293E/2020\|EPI_ISL_529630\|202 | ···················· | ·A·················· | ···················· | ···················· | ···················· | ···A················ |
| G | 8672 | hCoV-19/England/BIRM-6295C/2020\|EPI_ISL_529631\|202 | ···················· | ·A·················· | ···················· | ···················· | ···················· | ···A················ |
| G | 8673 | hCoV-19/England/BIRM-6296B/2020\|EPI_ISL_529632\|202 | ···················· | ···················· | ···················· | ···················· | ···················· | ···················· |
| G | 8674 | hCoV-19/England/BIRM-6297A/2020\|EPI_ISL_529633\|202 | ···················· | ·A·················· | ···················· | ···················· | ···················· | ···A················ |
| G | 8675 | hCoV-19/England/BIRM-62998/2020\|EPI_ISL_529634\|202 | ···················· | ·A·················· | ···················· | ···················· | ···················· | ···A················ |
| G | 8676 | hCoV-19/England/BIRM-629C5/2020\|EPI_ISL_529635\|202 | ···················· | ···················· | ···················· | ···················· | ···················· | ···················· |
| G | 8677 | hCoV-19/England/BIRM-629E3/2020\|EPI_ISL_529636\|202 | ···················· | ·A·················· | ···················· | ···················· | ···················· | ···A················ |
| G | 8678 | hCoV-19/England/BIRM-62A3B/2020\|EPI_ISL_529637\|202 | ···················· | ·A·················· | ···················· | ···················· | ···················· | ···A················ |
| G | 8679 | hCoV-19/England/BIRM-62A86/2020\|EPI_ISL_529638\|202 | ···················· | ·A·················· | ···················· | ···················· | ···················· | ···A················ |
| G | 8680 | hCoV-19/England/BIRM-62AA4/2020\|EPI_ISL_529639\|202 | ···················· | ·A·················· | ···················· | ···················· | ···················· | ···A················ |
| G | 8681 | hCoV-19/England/BIRM-62AB3/2020\|EPI_ISL_529640\|202 | ···················· | ···················· | ···················· | ···················· | ···················· | ···················· |
| G | 8682 | hCoV-19/England/BIRM-62AC2/2020\|EPI_ISL_529641\|202 | ···················· | ···················· | ···················· | ···················· | ···················· | ···················· |
| G | 8683 | hCoV-19/England/BIRM-62AD1/2020\|EPI_ISL_529642\|202 | ···················· | ···················· | ···················· | ···················· | ···················· | ···················· |

| | | | | | | | | |
|---|---|---|---|---|---|---|---|---|
| G | 8684 | hCoV-19/England/BIRM-62AE0/2020\|EPI_ISL_529643\|202 | ···················· | ·A·················· | ···················· | ···················· | ···················· | ··A················· |
| G | 8685 | hCoV-19/England/BIRM-63584/2020\|EPI_ISL_529644\|202 | ···················· | ···················· | ···················· | ···················· | ···················· | ···················· |
| G | 8686 | hCoV-19/England/BIRM-63706/2020\|EPI_ISL_529645\|202 | ···················· | ···················· | ···················· | ···················· | ···················· | ···················· |
| G | 8687 | hCoV-19/England/BIRM-63760/2020\|EPI_ISL_529646\|202 | ···················· | ·A·················· | ···················· | ···················· | ···················· | ···················· |
| G | 8688 | hCoV-19/England/BIRM-6377F/2020\|EPI_ISL_529647\|202 | ···················· | ·A·················· | ···················· | ···················· | ···················· | ···················· |
| G | 8689 | hCoV-19/England/BIRM-6378E/2020\|EPI_ISL_529648\|202 | ···················· | ·A·················· | ···················· | ···················· | ···················· | ···················· |
| G | 8690 | hCoV-19/England/BIRM-637CA/2020\|EPI_ISL_529649\|202 | ···················· | ···················· | ···················· | ···················· | ···················· | ···················· |
| G | 8691 | hCoV-19/England/BRIS-133C5A/2020\|EPI_ISL_529655\|20 | ···················· | ···················· | ···················· | ···················· | ···················· | ···················· |
| G | 8692 | hCoV-19/England/CAMB-1B5BBD/2020\|EPI_ISL_529656\|20 | ···················· | ···················· | ···················· | ···················· | ···················· | ···················· |
| G | 8693 | hCoV-19/England/CAMB-1B5C14/2020\|EPI_ISL_529657\|20 | ···················· | ···················· | ···················· | ···················· | ···················· | ···················· |
| G | 8694 | hCoV-19/England/CAMB-1B5C23/2020\|EPI_ISL_529658\|20 | ···················· | ···················· | ···················· | ···················· | ···················· | ···················· |
| G | 8695 | hCoV-19/England/CAMB-1B5C32/2020\|EPI_ISL_529659\|20 | ···················· | ···················· | ···················· | ···················· | ···················· | ···················· |
| G | 8696 | hCoV-19/Scotland/EDB6845/2020\|EPI_ISL_529691\|2020- | ···················· | ···················· | ···················· | ···················· | ···················· | ···················· |
| G | 8697 | hCoV-19/Scotland/EDB6935/2020\|EPI_ISL_529692\|2020- | ···················· | ···················· | ···················· | ···················· | ···················· | ···················· |
| G | 8698 | hCoV-19/Scotland/EDB7366/2020\|EPI_ISL_529693\|2020- | ···················· | ···················· | ···················· | ···················· | ···················· | ···················· |
| G | 8699 | hCoV-19/Scotland/EDB7373/2020\|EPI_ISL_529694\|2020- | ···················· | ···················· | ···················· | ···················· | ···················· | ···················· |
| G | 8700 | hCoV-19/Scotland/EDB7374/2020\|EPI_ISL_529695\|2020- | ···················· | ···················· | ···················· | ···················· | ···················· | ···················· |
| G | 8701 | hCoV-19/Scotland/EDB7378/2020\|EPI_ISL_529696\|2020- | ···················· | ···················· | ···················· | ···················· | ···················· | ···················· |
| G | 8703 | hCoV-19/Wales/PHWC-169C88/2020\|EPI_ISL_529698\|2020 | ···················· | ···················· | ···················· | ···················· | ···················· | ···················· |
| G | 8704 | hCoV-19/Wales/PHWC-169CA6/2020\|EPI_ISL_529699\|2020 | ···················· | ···················· | ···················· | ···················· | ···················· | ···················· |
| G | 8705 | hCoV-19/Peru/LAL-INS-056/2020\|EPI_ISL_517686\|2020- | ···················· | ···················· | ···················· | ···················· | ···················· | ···················· |
| G | 8706 | hCoV-19/Peru/ANC-INS-059/2020\|EPI_ISL_517687\|2020- | ···················· | ···················· | ···················· | ···················· | ···················· | ···················· |
| G | 8707 | hCoV-19/Suriname/SR-52/2020\|EPI_ISL_518801\|2020-07 | ···············A···· | ···················· | ···················· | ···················· | ···················· | ···················· |
| G | 8708 | hCoV-19/Suriname/SR-53/2020\|EPI_ISL_518802\|2020-07 | ···················· | ···················· | ···················· | ···················· | ···················· | ···················· |
| G | 8709 | hCoV-19/Suriname/SR-54/2020\|EPI_ISL_518803\|2020-07 | ···············A···· | ···················· | ···················· | ···················· | ···················· | ···················· |
| G | 8710 | hCoV-19/Suriname/SR-55/2020\|EPI_ISL_518804\|2020-07 | ···················· | ···················· | ···················· | ···················· | ···················· | ···················· |
| G | 8711 | hCoV-19/Suriname/SR-56/2020\|EPI_ISL_518805\|2020-07 | ···················· | ···················· | ···················· | ···················· | ···················· | ···················· |
| G | 8713 | hCoV-19/Australia/VIC2330/2020\|EPI_ISL_522051\|2020 | ···················· | ···················· | ···················· | ···················· | ···················· | ···················· |
| G | 8715 | hCoV-19/Australia/VIC2332/2020\|EPI_ISL_522053\|2020 | ···················· | ···················· | ···················· | ···················· | ···················· | ···················· |
| G | 8716 | hCoV-19/Australia/VIC2340/2020\|EPI_ISL_522061\|2020 | ···················· | ···················· | ···················· | ···················· | ···················· | ···················· |
| G | 8717 | hCoV-19/Australia/VIC2341/2020\|EPI_ISL_522062\|2020 | ···················· | ···················· | ···················· | ···················· | ···················· | ···················· |
| G | 8718 | hCoV-19/Australia/VIC2342/2020\|EPI_ISL_522063\|2020 | ···················· | ···················· | ···················· | ···················· | ···················· | ···················· |
| G | 8719 | hCoV-19/Australia/VIC2343/2020\|EPI_ISL_522064\|2020 | ···················· | ···················· | ···················· | ···················· | ···················· | ···················· |
| G | 8720 | hCoV-19/Australia/VIC2344/2020\|EPI_ISL_522065\|2020 | ···················· | ···················· | ···················· | ···················· | ···················· | ···················· |
| G | 8721 | hCoV-19/Australia/VIC2345/2020\|EPI_ISL_522066\|2020 | ···················· | ···················· | ···················· | ···················· | ···················· | ···················· |
| G | 8722 | hCoV-19/Australia/VIC2346/2020\|EPI_ISL_522067\|2020 | ···················· | ···················· | ···················· | ···················· | ···················· | ···················· |
| G | 8723 | hCoV-19/Australia/VIC2347/2020\|EPI_ISL_522068\|2020 | ···················· | ···················· | ···················· | ···················· | ···················· | ···················· |
| G | 8724 | hCoV-19/Australia/VIC2349/2020\|EPI_ISL_522069\|2020 | ···················· | ···················· | ···················· | ···················· | ···················· | ···················· |
| G | 8725 | hCoV-19/Netherlands/NB-EMC-178/2020\|EPI_ISL_523301 | ···················· | ···················· | ···················· | ···················· | ···················· | ···················· |
| G | 8726 | hCoV-19/Netherlands/NB-EMC-180/2020\|EPI_ISL_523303 | ···················· | ···················· | ···················· | ···················· | ···················· | ···················· |
| G | 8727 | hCoV-19/Netherlands/NB-EMC-181/2020\|EPI_ISL_523304 | ···················· | ···················· | ···················· | ···················· | ···················· | ···················· |
| G | 8728 | hCoV-19/Netherlands/NB-EMC-182/2020\|EPI_ISL_523305 | ···················· | ···················· | ···················· | ···················· | ···················· | ···················· |
| G | 8729 | hCoV-19/Netherlands/NB-EMC-183/2020\|EPI_ISL_523306 | ···················· | ···················· | ···················· | ···················· | ···················· | ···················· |
| G | 8730 | hCoV-19/Netherlands/NB-EMC-184/2020\|EPI_ISL_523307 | ···················· | ···················· | ···················· | ···················· | ···················· | ···················· |
| G | 8731 | hCoV-19/Netherlands/NB-EMC-185/2020\|EPI_ISL_523308 | ···················· | ···················· | ···················· | ···················· | ···················· | ···················· |
| G | 8732 | hCoV-19/Netherlands/NB-EMC-186/2020\|EPI_ISL_523309 | ···················· | ···················· | ···················· | ···················· | ···················· | ···················· |
| G | 8733 | hCoV-19/Netherlands/NB-EMC-187/2020\|EPI_ISL_523310 | ···················· | ···················· | ···················· | ···················· | ···················· | ···················· |
| G | 8734 | hCoV-19/Netherlands/NB-EMC-188/2020\|EPI_ISL_523311 | ···················· | ···················· | ···················· | ···················· | ···················· | ···················· |
| G | 8735 | hCoV-19/Netherlands/NB-EMC-189/2020\|EPI_ISL_523312 | ···················· | ···················· | ···················· | ···················· | ···················· | ···················· |
| G | 8736 | hCoV-19/Netherlands/NB-EMC-190/2020\|EPI_ISL_523313 | ···················· | ···················· | ···················· | ···················· | ···················· | ···················· |
| G | 8737 | hCoV-19/Netherlands/NB-EMC-191/2020\|EPI_ISL_523314 | ···················· | ···················· | ···················· | ···················· | ···················· | ···················· |
| G | 8738 | hCoV-19/Netherlands/NB-EMC-192/2020\|EPI_ISL_523315 | ···················· | ···················· | ···················· | ···················· | ···················· | ···················· |
| G | 8739 | hCoV-19/Netherlands/NB-EMC-257/2020\|EPI_ISL_523380 | ···················· | ···················· | ···················· | ···················· | ·····T·············· | ···················· |
| G | 8740 | hCoV-19/Netherlands/NB-EMC-258/2020\|EPI_ISL_523381 | ···················· | ···················· | ···················· | ···················· | ·····T·············· | ···················· |
| G | 8741 | hCoV-19/Netherlands/NB-EMC-259/2020\|EPI_ISL_523382 | ···················· | ···················· | ···················· | ···················· | ···················· | ···················· |
| G | 8742 | hCoV-19/Netherlands/NB-EMC-260/2020\|EPI_ISL_523383 | ···················· | ···················· | ···················· | ···················· | ···················· | ···················· |
| G | 8743 | hCoV-19/Netherlands/NB-EMC-261/2020\|EPI_ISL_523384 | ···················· | ···················· | ···················· | ···················· | ···················· | ···················· |
| G | 8744 | hCoV-19/Netherlands/NB-EMC-272/2020\|EPI_ISL_523394 | ···················· | ···················· | ···················· | ···················· | ···················· | ···················· |
| G | 8745 | hCoV-19/Netherlands/NB-EMC-273/2020\|EPI_ISL_523395 | ···················· | ·····A·············· | ···················· | ···················· | ···················· | ···················· |
| G | 8746 | hCoV-19/Netherlands/NB-EMC-274/2020\|EPI_ISL_523396 | ···················· | ·····A·············· | ···················· | ···················· | ···················· | ···················· |
| G | 8747 | hCoV-19/Netherlands/NB-EMC-275/2020\|EPI_ISL_523397 | ···················· | ·A·················· | ···················· | ···················· | ···················· | ··A················· |
| G | 8748 | hCoV-19/England/CAMB-1AE267/2020\|EPI_ISL_524600\|20 | ···················· | ···················· | ···················· | ···················· | ···················· | ···················· |
| G | 8749 | hCoV-19/England/CAMB-1AE285/2020\|EPI_ISL_524601\|20 | ···················· | ···················· | ···················· | ···················· | ···················· | ···················· |
| G | 8750 | hCoV-19/England/CAMB-1AEA50/2020\|EPI_ISL_524603\|20 | ···················· | ···················· | ···················· | ···················· | ···················· | ···················· |
| G | 8751 | hCoV-19/England/CAMB-1AE944/2020\|EPI_ISL_524604\|20 | ···················· | ···················· | ···················· | ···················· | ···················· | ···················· |
| G | 8752 | hCoV-19/England/CAMB-1AE74A/2020\|EPI_ISL_524606\|20 | ···················· | ···················· | ···················· | ···················· | ···················· | ···················· |
| G | 8753 | hCoV-19/England/CAMB-1AE0F4/2020\|EPI_ISL_524608\|20 | ···················· | ···················· | ···················· | ···················· | ···················· | ···················· |
| G | 8754 | hCoV-19/England/CAMB-1AE838/2020\|EPI_ISL_524609\|20 | ···················· | ···················· | ···················· | ···················· | ···················· | ···················· |
| G | 8755 | hCoV-19/England/CAMB-1AEB20/2020\|EPI_ISL_524610\|20 | ···················· | ···················· | ···················· | ···················· | ···················· | ···················· |
| G | 8756 | hCoV-19/England/CAMB-1AE865/2020\|EPI_ISL_524612\|20 | ···················· | ···················· | ···················· | ···················· | ···················· | ···················· |
| G | 8757 | hCoV-19/England/CAMB-1AE7B3/2020\|EPI_ISL_524615\|20 | ···················· | ···················· | ···················· | ···················· | ···················· | ···················· |
| G | 8758 | hCoV-19/England/CAMB-1AE62F/2020\|EPI_ISL_524616\|20 | ···················· | ···················· | ···················· | ···················· | ···················· | ···················· |
| G | 8759 | hCoV-19/England/CAMB-1AE73B/2020\|EPI_ISL_524617\|20 | ···················· | ···················· | ···················· | ···················· | ···················· | ···················· |
| G | 8760 | hCoV-19/England/CAMB-1AE7FF/2020\|EPI_ISL_524618\|20 | ···················· | ···················· | ···················· | ···················· | ···················· | ···················· |
| G | 8761 | hCoV-19/England/LOND-126118D/2020\|EPI_ISL_524619\|2 | ···················· | ···················· | ···················· | ···················· | ···················· | ···················· |
| G | 8762 | hCoV-19/England/LOND-12612C6/2020\|EPI_ISL_524620\|2 | ···················· | ···················· | ···················· | ···················· | ···················· | ···················· |
| G | 8763 | hCoV-19/England/LOND-1261211/2020\|EPI_ISL_524621\|2 | ···················· | ···················· | ···················· | ···················· | ···················· | ···················· |
| G | 8764 | hCoV-19/England/LOND-126027C/2020\|EPI_ISL_524622\|2 | ···················· | ···················· | ···················· | ···················· | ···················· | ···················· |
| G | 8765 | hCoV-19/England/LOND-126133C/2020\|EPI_ISL_524623\|2 | ···················· | ···················· | ···················· | ···················· | ···················· | ···················· |
| G | 8766 | hCoV-19/England/LOND-12611BA/2020\|EPI_ISL_524624\|2 | ···················· | ····A··············· | ···················· | ···················· | ···················· | ···················· |

| | | | | | | | | |
|---|---|---|---|---|---|---|---|---|
| G | 8767 | hCoV-19/England/LOND-126131E/2020\|EPI_ISL_524625\|2 | ···················· | ·············A······ | ····················· | ···················· | ···················· | ···················· |
| G | 8768 | hCoV-19/England/LOND-126124E/2020\|EPI_ISL_524628\|2 | ···················· | ···················· | ····················· | ···················· | ···················· | ···················· |
| G | 8769 | hCoV-19/England/LOND-1261080/2020\|EPI_ISL_524629\|2 | ···················· | ···················· | ····················· | ···················· | ···················· | ···················· |
| G | 8770 | hCoV-19/England/LOND-126109F/2020\|EPI_ISL_524630\|2 | ···················· | ···················· | ····················· | ···················· | ···················· | ···················· |
| G | 8771 | hCoV-19/England/LOND-12611E7/2020\|EPI_ISL_524631\|2 | ···················· | ···················· | ····················· | ···················· | ···················· | ···················· |
| G | 8772 | hCoV-19/England/LOND-12612F3/2020\|EPI_ISL_524632\|2 | ···················· | ···················· | ····················· | ···················· | ···················· | ···················· |
| G | 8773 | hCoV-19/England/LOND-1261114/2020\|EPI_ISL_524633\|2 | ···················· | ···················· | ····················· | ···················· | ···················· | ···················· |
| G | 8774 | hCoV-19/England/LOND-1261105/2020\|EPI_ISL_524634\|2 | ···················· | ···················· | ····················· | ···················· | ···················· | ···················· |
| G | 8775 | hCoV-19/England/LOND-1261299/2020\|EPI_ISL_524636\|2 | ···················· | ···················· | ····················· | ···················· | ···················· | ···················· |
| G | 8776 | hCoV-19/England/LOND-1260C6F/2020\|EPI_ISL_524638\|2 | ···················· | ···················· | ····················· | ···················· | ···················· | ···················· |
| G | 8777 | hCoV-19/England/LOND-126085C/2020\|EPI_ISL_524640\|2 | ···················· | ···················· | ····················· | ···················· | ···················· | ···················· |
| G | 8778 | hCoV-19/England/LOND-12604EF/2020\|EPI_ISL_524641\|2 | ···················· | ···················· | ····················· | ···················· | ···················· | ···················· |
| G | 8779 | hCoV-19/England/LOND-1260A74/2020\|EPI_ISL_524642\|2 | ···················· | ···················· | ····················· | ···················· | ···················· | ···················· |
| G | 8780 | hCoV-19/England/LOND-1260E4B/2020\|EPI_ISL_524643\|2 | ···················· | ···················· | ····················· | ···················· | ···················· | ···················· |
| G | 8781 | hCoV-19/England/LOND-1260C41/2020\|EPI_ISL_524645\|2 | ···················· | ···················· | ····················· | ···················· | ···················· | ···················· |
| G | 8782 | hCoV-19/England/LOND-1260B17/2020\|EPI_ISL_524646\|2 | ···················· | ···················· | ····················· | ···················· | ···················· | ···················· |
| G | 8783 | hCoV-19/USA/OR-OHSU-0785/2020\|EPI_ISL_525900\|2020- | ···················· | ···················· | ····················· | ···················· | ···················· | ···················· |
| G | 8784 | hCoV-19/USA/OR-OHSU-0797/2020\|EPI_ISL_525901\|2020- | ···················· | ···················· | ····················· | ···················· | ···················· | ···················· |
| G | 8785 | hCoV-19/USA/OR-OHSU-0801/2020\|EPI_ISL_525902\|2020- | ···················· | ·A·················· | ····················· | ···················· | ···················· | ···················· |
| G | 8786 | hCoV-19/USA/OR-OHSU-0803/2020\|EPI_ISL_525903\|2020- | ···················· | ·A·················· | ····················· | ···················· | ···················· | ···················· |
| G | 8787 | hCoV-19/USA/OR-OHSU-0807/2020\|EPI_ISL_525904\|2020- | ···················· | ···················· | ····················· | ···················· | ···················· | ···················· |
| G | 8788 | hCoV-19/USA/OR-OHSU-0809/2020\|EPI_ISL_525905\|2020- | ···················· | ···················· | ····················· | ···················· | ···················· | ···················· |
| G | 8789 | hCoV-19/USA/OR-OHSU-0811/2020\|EPI_ISL_525906\|2020- | ···················· | ···················· | ····················· | ···················· | ···················· | ···················· |
| G | 8790 | hCoV-19/USA/OR-OHSU-0817/2020\|EPI_ISL_525907\|2020- | ···················· | ···················· | ····················· | ···················· | ···················· | ···················· |
| G | 8791 | hCoV-19/USA/OR-OHSU-0819/2020\|EPI_ISL_525908\|2020- | ···················· | ···················· | ····················· | ···················· | ···················· | ···················· |
| G | 8792 | hCoV-19/USA/OR-OHSU-0827/2020\|EPI_ISL_525909\|2020- | ···················· | ···················· | ····················· | ···················· | ···················· | ···················· |
| G | 8793 | hCoV-19/USA/OR-OHSU-0829/2020\|EPI_ISL_525910\|2020- | ···················· | ···················· | ····················· | ···················· | ···················· | ···················· |
| G | 8794 | hCoV-19/USA/OR-OHSU-0851/2020\|EPI_ISL_525911\|2020- | ···················· | ···················· | ····················· | ···················· | ···················· | ···················· |
| G | 8795 | hCoV-19/USA/OR-OHSU-0855/2020\|EPI_ISL_525912\|2020- | ···················· | ···················· | ····················· | ···················· | ···················· | ···················· |
| G | 8796 | hCoV-19/USA/OR-OHSU-0859/2020\|EPI_ISL_525913\|2020- | ···················· | ···················· | ····················· | ···················· | ···················· | ···················· |
| G | 8797 | hCoV-19/USA/OR-OHSU-0865/2020\|EPI_ISL_525914\|2020- | ···················· | ···················· | ····················· | ···················· | ···················· | ···················· |
| G | 8798 | hCoV-19/USA/OR-OHSU-0867/2020\|EPI_ISL_525915\|2020- | ···················· | ···················· | ····················· | ···················· | ···················· | ···················· |
| G | 8799 | hCoV-19/USA/OR-OHSU-0869/2020\|EPI_ISL_525916\|2020- | ···················· | ···················· | ····················· | ···················· | ···················· | ···················· |
| G | 8800 | hCoV-19/USA/OR-OHSU-0871/2020\|EPI_ISL_525917\|2020- | ···················· | ···················· | ····················· | ···················· | ···················· | ···················· |
| G | 8801 | hCoV-19/USA/OR-OHSU-0875/2020\|EPI_ISL_525918\|2020- | ···················· | ···················· | ····················· | ···················· | ···················· | ···················· |
| G | 8802 | hCoV-19/USA/OR-OHSU-0877/2020\|EPI_ISL_525919\|2020- | ···················· | ···················· | ····················· | ···················· | ···················· | ···················· |
| G | 8803 | hCoV-19/USA/OR-OHSU-0879/2020\|EPI_ISL_525920\|2020- | ···················· | ·A·················· | ····················· | ···················· | ···················· | ···················· |
| G | 8804 | hCoV-19/USA/OR-OHSU-0881/2020\|EPI_ISL_525921\|2020- | ···················· | ···················· | ····················· | ···················· | ···················· | ···················· |
| G | 8805 | hCoV-19/USA/OR-OHSU-0899/2020\|EPI_ISL_525922\|2020- | ···················· | ···················· | ····················· | ···················· | ···················· | ···················· |
| G | 8806 | hCoV-19/USA/OR-OHSU-0901/2020\|EPI_ISL_525923\|2020- | ···················· | ···················· | ····················· | ···················· | ···················· | ···················· |
| G | 8807 | hCoV-19/USA/OR-OHSU-0905/2020\|EPI_ISL_525924\|2020- | ···················· | ···················· | ····················· | ···················· | ···················· | ···················· |
| G | 8808 | hCoV-19/USA/OR-OHSU-0911/2020\|EPI_ISL_525925\|2020- | ···················· | ·A·················· | ····················· | ···················· | ···················· | ···················· |
| G | 8809 | hCoV-19/USA/OR-OHSU-0919/2020\|EPI_ISL_525927\|2020- | ···················· | ···················· | ····················· | ···················· | ···················· | ···················· |
| G | 8810 | hCoV-19/USA/OR-OHSU-0921/2020\|EPI_ISL_525928\|2020- | ···················· | ···················· | ····················· | ···················· | ···················· | ···················· |
| G | 8811 | hCoV-19/USA/OR-OHSU-0923/2020\|EPI_ISL_525929\|2020- | ···················· | ···················· | ····················· | ···················· | ···················· | ···················· |
| G | 8812 | hCoV-19/USA/OR-OHSU-0929/2020\|EPI_ISL_525930\|2020- | ···················· | ···················· | ····················· | ···················· | ···················· | ···················· |
| G | 8813 | hCoV-19/USA/OR-OHSU-0937/2020\|EPI_ISL_525931\|2020- | ···················· | ·A·················· | ····················· | ···················· | ···················· | ···················· |
| G | 8814 | hCoV-19/USA/OR-OHSU-0941/2020\|EPI_ISL_525932\|2020- | ···················· | ···················· | ····················· | ···················· | ···················· | ···················· |
| G | 8815 | hCoV-19/USA/OR-OHSU-0945/2020\|EPI_ISL_525933\|2020- | ···················· | ···················· | ····················· | ···················· | ···················· | ···················· |
| G | 8816 | hCoV-19/USA/OR-OHSU-0951/2020\|EPI_ISL_525934\|2020- | ···················· | ···················· | ····················· | ···················· | ···················· | ···················· |
| G | 8817 | hCoV-19/USA/OR-OHSU-0953/2020\|EPI_ISL_525935\|2020- | ···················· | ···················· | ····················· | ···················· | ···················· | ···················· |
| G | 8818 | hCoV-19/USA/OR-OHSU-0959/2020\|EPI_ISL_525936\|2020- | ···················· | ···················· | ····················· | ···················· | ···················· | ···················· |
| G | 8819 | hCoV-19/USA/OR-OHSU-0963/2020\|EPI_ISL_525937\|2020- | ···················· | ···················· | ····················· | ···················· | ···················· | ···················· |
| G | 8820 | hCoV-19/USA/OR-OHSU-0967/2020\|EPI_ISL_525938\|2020- | ···················· | ·A·················· | ····················· | ···················· | ···················· | ···················· |
| G | 8821 | hCoV-19/USA/OR-OHSU-0969/2020\|EPI_ISL_525939\|2020- | ···················· | ·A·················· | ····················· | ···················· | ···················· | ···················· |
| G | 8822 | hCoV-19/USA/OR-OHSU-0979/2020\|EPI_ISL_525940\|2020- | ···················· | ···················· | ····················· | ···················· | ···················· | ···················· |
| G | 8823 | hCoV-19/USA/OR-OHSU-0981/2020\|EPI_ISL_525941\|2020- | ···················· | ···················· | ····················· | ···················· | ···················· | ···················· |
| G | 8824 | hCoV-19/USA/OR-OHSU-0983/2020\|EPI_ISL_525942\|2020- | ···················· | ···················· | ····················· | ···················· | ···················· | ···················· |
| G | 8825 | hCoV-19/USA/OR-OHSU-0985/2020\|EPI_ISL_525943\|2020- | ···················· | ·A·················· | ····················· | ···················· | ···················· | ···················· |
| G | 8826 | hCoV-19/USA/OR-OHSU-0993/2020\|EPI_ISL_525944\|2020- | ···················· | ·A·················· | ····················· | ···················· | ···················· | ···················· |
| G | 8827 | hCoV-19/USA/OR-OHSU-1003/2020\|EPI_ISL_525945\|2020- | ···················· | ···················· | ····················· | ···················· | ···················· | ···················· |
| G | 8828 | hCoV-19/USA/OR-OHSU-1013/2020\|EPI_ISL_525947\|2020- | ···················· | ·A·················· | ····················· | ···················· | ···················· | ···················· |
| G | 8829 | hCoV-19/USA/OR-OHSU-1015/2020\|EPI_ISL_525948\|2020- | ···················· | ·A·················· | ····················· | ···················· | ···················· | ···················· |
| G | 8830 | hCoV-19/USA/OR-OHSU-1019/2020\|EPI_ISL_525949\|2020- | ···················· | ·············A······ | ····················· | ···················· | ···················· | ···················· |
| G | 8831 | hCoV-19/USA/OR-OHSU-1021/2020\|EPI_ISL_525950\|2020- | ···················· | ···················· | ····················· | ···················· | ···················· | ···················· |
| G | 8832 | hCoV-19/USA/OR-OHSU-1023/2020\|EPI_ISL_525951\|2020- | ···················· | ·A·················· | ····················· | ···················· | ···················· | ···················· |
| G | 8833 | hCoV-19/USA/OR-OHSU-1027/2020\|EPI_ISL_525952\|2020- | ···················· | ·A·················· | ····················· | ···················· | ···················· | ···················· |
| G | 8834 | hCoV-19/USA/OR-OHSU-1029/2020\|EPI_ISL_525953\|2020- | ···················· | ·A·················· | ····················· | ···················· | ···················· | ···················· |
| G | 8835 | hCoV-19/USA/OR-OHSU-1031/2020\|EPI_ISL_525954\|2020- | ···················· | ···················· | ····················· | ···················· | ···················· | ···················· |
| G | 8836 | hCoV-19/USA/OR-OHSU-1035/2020\|EPI_ISL_525955\|2020- | ···················· | ·A·················· | ····················· | ···················· | ···················· | ···················· |
| G | 8837 | hCoV-19/USA/OR-OHSU-1041/2020\|EPI_ISL_525956\|2020- | ···················· | ···················· | ····················· | ···················· | ···················· | ···················· |
| G | 8838 | hCoV-19/USA/OR-OHSU-1043/2020\|EPI_ISL_525957\|2020- | ···················· | ···················· | ····················· | ···················· | ···················· | ···················· |
| G | 8839 | hCoV-19/USA/OR-OHSU-1047/2020\|EPI_ISL_525958\|2020- | ···················· | ·A·················· | ····················· | ···················· | ···················· | ···················· |
| G | 8840 | hCoV-19/USA/OR-OHSU-1049/2020\|EPI_ISL_525959\|2020- | ···················· | ···················· | ····················· | ···················· | ···················· | ···················· |
| G | 8841 | hCoV-19/USA/OR-OHSU-1059/2020\|EPI_ISL_525960\|2020- | ···················· | ···················· | ····················· | ···················· | ···················· | ···················· |
| G | 8842 | hCoV-19/USA/OR-OHSU-1065/2020\|EPI_ISL_525961\|2020- | ···················· | ·A·················· | ····················· | ···················· | ···················· | ···················· |
| G | 8843 | hCoV-19/USA/OR-OHSU-1069/2020\|EPI_ISL_525962\|2020- | ···················· | ·A·················· | ····················· | ···················· | ···················· | ···················· |
| G | 8844 | hCoV-19/USA/OR-OHSU-1071/2020\|EPI_ISL_525963\|2020- | ···················· | ·A·················· | ····················· | ···················· | ···················· | ···················· |
| G | 8845 | hCoV-19/USA/OR-OHSU-1075/2020\|EPI_ISL_525964\|2020- | ···················· | ·A·················· | ····················· | ···················· | ···················· | ···················· |
| G | 8846 | hCoV-19/USA/OR-OHSU-1079/2020\|EPI_ISL_525965\|2020- | ···················· | ···················· | ····················· | ···················· | ···················· | ···················· |

| | | | | | | | |
|---|---|---|---|---|---|---|---|
| G | 8847 hCoV-19/USA/OR-OHSU-1083/2020\|EPI_ISL_525966\|2020- | ···················· | ·A·················· | ···················· | ···················· | ···················· | ···················· |
| G | 8848 hCoV-19/USA/OR-OHSU-1085/2020\|EPI_ISL_525967\|2020- | ···················· | ···················· | ···················· | ···················· | ···················· | ···················· |
| G | 8849 hCoV-19/USA/OR-OHSU-1101/2020\|EPI_ISL_525969\|2020- | ···················· | ···················· | ···················· | ···················· | ···················· | ···················· |
| G | 8850 hCoV-19/USA/OR-OHSU-1103/2020\|EPI_ISL_525970\|2020- | ···················· | ···················· | ···················· | ···················· | ···················· | ···················· |
| G | 8851 hCoV-19/USA/OR-OHSU-1489/2020\|EPI_ISL_525971\|2020- | ···················· | ···················· | ···················· | ···················· | ···················· | ···················· |
| G | 8852 hCoV-19/USA/OR-OHSU-1491/2020\|EPI_ISL_525972\|2020- | ···················· | ·A·················· | ···················· | ···················· | ···················· | ···················· |
| G | 8853 hCoV-19/USA/OR-OHSU-1493/2020\|EPI_ISL_525973\|2020- | ···················· | ···················· | ···················· | ···················· | ···················· | ···················· |
| G | 8854 hCoV-19/USA/OR-OHSU-1495/2020\|EPI_ISL_525974\|2020- | ···················· | ·A·················· | ···················· | ···················· | ···················· | ···················· |
| G | 8855 hCoV-19/USA/OR-OHSU-1535/2020\|EPI_ISL_525975\|2020- | ···················· | ···················· | ···················· | ···················· | ···················· | ···················· |
| G | 8856 hCoV-19/USA/OR-OHSU-1537/2020\|EPI_ISL_525976\|2020- | ···················· | ···················· | ···················· | ···················· | ···················· | ···················· |
| G | 8857 hCoV-19/USA/OR-OHSU-1551/2020\|EPI_ISL_525977\|2020- | ···················· | ···················· | ···················· | ···················· | ···················· | ···················· |
| G | 8858 hCoV-19/Egypt/EGY-043/2020\|EPI_ISL_527000\|2020-08- | ···················· | ···················· | ···················· | ···················· | ···················· | ···················· |
| G | 8859 hCoV-19/Egypt/EGY-045/2020\|EPI_ISL_527001\|2020-08- | ···················· | ···················· | ···················· | ···················· | ···················· | ···················· |
| G | 8860 hCoV-19/Egypt/EGY-046/2020\|EPI_ISL_527002\|2020-08- | ···················· | ···················· | ···················· | ···················· | ···················· | ···················· |
| G | 8861 hCoV-19/Egypt/EGY-047/2020\|EPI_ISL_527003\|2020-08- | ···················· | ···················· | ···················· | ···················· | ···················· | ···················· |
| G | 8862 hCoV-19/Egypt/EGY-048/2020\|EPI_ISL_527004\|2020-08- | ···················· | ···················· | ···················· | ···················· | ···················· | ···················· |
| G | 8863 hCoV-19/Egypt/EGY-049/2020\|EPI_ISL_527005\|2020-08- | ···················· | ···················· | ···················· | ···················· | ···················· | ···················· |
| G | 8864 hCoV-19/Egypt/EGY-050/2020\|EPI_ISL_527006\|2020-08- | ···················· | ···················· | ···················· | ···················· | ···················· | ···················· |
| G | 8865 hCoV-19/Egypt/EGY-040/2020\|EPI_ISL_527007\|2020-08- | ···················· | ···················· | ···················· | ···················· | ···················· | ···················· |
| G | 8866 hCoV-19/Australia/NSW2144/2020\|EPI_ISL_527008\|2020 | ···················· | ···················· | ···················· | ···················· | ···················· | ···················· |
| G | 8867 hCoV-19/Australia/NSW2193/2020\|EPI_ISL_527009\|2020 | ···················· | ···················· | ···················· | ···················· | ···················· | ···················· |
| G | 8868 hCoV-19/Australia/NSW2254/2020\|EPI_ISL_527010\|2020 | ···················· | ···················· | ···················· | ···················· | ···················· | ···················· |
| G | 8869 hCoV-19/Australia/NSW2410/2020\|EPI_ISL_527011\|2020 | ···················· | ···················· | ···················· | ···················· | ···················· | ···················· |
| G | 8870 hCoV-19/Australia/NSW2411/2020\|EPI_ISL_527012\|2020 | ···················· | ···················· | ···················· | ···················· | ···················· | ···················· |
| G | 8871 hCoV-19/Australia/NSW2415/2020\|EPI_ISL_527013\|2020 | ···················· | ···················· | ···················· | ···················· | ···················· | ···················· |
| G | 8872 hCoV-19/Australia/NSW2429/2020\|EPI_ISL_527014\|2020 | ···················· | ···················· | ···················· | ···················· | ···················· | ···················· |
| G | 8873 hCoV-19/Australia/NSW2431/2020\|EPI_ISL_527015\|2020 | ···················· | ···················· | ···················· | ···················· | ···················· | ···················· |
| G | 8874 hCoV-19/Australia/NSW2435/2020\|EPI_ISL_527016\|2020 | ···················· | ···················· | ···················· | ···················· | ···················· | ···················· |
| G | 8875 hCoV-19/Australia/NSW2437/2020\|EPI_ISL_527017\|2020 | ···················· | ···················· | ···················· | ···················· | ···················· | ···················· |
| G | 8876 hCoV-19/Australia/NSW2456/2020\|EPI_ISL_527018\|2020 | ···················· | ···················· | ···················· | ···················· | ···················· | ···················· |
| G | 8877 hCoV-19/Australia/NSW2473/2020\|EPI_ISL_527019\|2020 | ···················· | ···················· | ···················· | ···················· | ···················· | ···················· |
| G | 8878 hCoV-19/Australia/NSW2474/2020\|EPI_ISL_527020\|2020 | ···················· | ···················· | ···················· | ···················· | ···················· | ···················· |
| G | 8879 hCoV-19/Australia/NSW2480/2020\|EPI_ISL_527021\|2020 | ···················· | ·A·················· | ···················· | ···················· | ···················· | ··A················· |
| G | 8880 hCoV-19/Australia/NSW2486/2020\|EPI_ISL_527022\|2020 | ···················· | ···················· | ···················· | ···················· | ···················· | ···················· |
| G | 8881 hCoV-19/Australia/NSW2487/2020\|EPI_ISL_527023\|2020 | ···················· | ···················· | ···················· | ···················· | ···················· | ···················· |
| G | 8882 hCoV-19/Australia/NSW2495/2020\|EPI_ISL_527024\|2020 | ···················· | ···················· | ···················· | ···················· | ···················· | ···················· |
| G | 8883 hCoV-19/Australia/NSW2496/2020\|EPI_ISL_527025\|2020 | ···················· | ···················· | ···················· | ···················· | ···················· | ···················· |
| G | 8884 hCoV-19/Australia/NSW2497/2020\|EPI_ISL_527026\|2020 | ···················· | ···················· | ···················· | ···················· | ···················· | ···················· |
| G | 8885 hCoV-19/Australia/NSW2503/2020\|EPI_ISL_527027\|2020 | ···················· | ···················· | ···················· | ···················· | ···················· | ···················· |
| G | 8886 hCoV-19/Australia/NSW2507/2020\|EPI_ISL_527028\|2020 | ···················· | ···················· | ···················· | ···················· | ···················· | ···················· |
| G | 8887 hCoV-19/Australia/NSW2508/2020\|EPI_ISL_527029\|2020 | ···················· | ···················· | ···················· | ···················· | ···················· | ···················· |
| G | 8888 hCoV-19/Australia/NSW2536/2020\|EPI_ISL_527039\|2020 | ···················· | ···················· | ···················· | ···················· | ···················· | ···················· |
| G | 8889 hCoV-19/Switzerland/AG-UHB-42195061/2020\|EPI_ISL_5 | ···················· | ···················· | ···················· | ···················· | ···················· | ···················· |
| G | 8890 hCoV-19/Switzerland/BL-UHB-42195083/2020\|EPI_ISL_5 | ···················· | ···················· | ···················· | ···················· | ···················· | ···················· |
| G | 8891 hCoV-19/Switzerland/SO-UHB-42195124/2020\|EPI_ISL_5 | ···················· | ···················· | ···················· | ···················· | ···················· | ···················· |
| G | 8892 hCoV-19/Switzerland/BS-UHB-42195295/2020\|EPI_ISL_5 | ···················· | ···················· | ···················· | ···················· | ···················· | ···················· |
| G | 8893 hCoV-19/Switzerland/BL-UHB-42195424/2020\|EPI_ISL_5 | ···················· | ···················· | ···················· | ···················· | ···················· | ···················· |
| G | 8894 hCoV-19/Switzerland/BS-UHB-42195425/2020\|EPI_ISL_5 | ···················· | ···················· | ···················· | ···················· | ···················· | ···················· |
| G | 8895 hCoV-19/Switzerland/BS-UHB-42195540/2020\|EPI_ISL_5 | ···················· | ···················· | ···················· | ···················· | ···················· | ···················· |
| G | 8896 hCoV-19/Switzerland/BL-UHB-42195605/2020\|EPI_ISL_5 | ···················· | ············A······· | ···················· | ···················· | ···················· | ···················· |
| G | 8897 hCoV-19/Switzerland/BL-UHB-42195622/2020\|EPI_ISL_5 | ···················· | ···················· | ···················· | ···················· | ···················· | ···················· |
| G | 8898 hCoV-19/Switzerland/BS-UHB-42195640/2020\|EPI_ISL_5 | ···················· | ···················· | ···················· | ···················· | ···················· | ···················· |
| G | 8899 hCoV-19/Switzerland/BS-UHB-42195693/2020\|EPI_ISL_5 | ···················· | ···················· | ···················· | ···················· | ···················· | ···················· |
| G | 8900 hCoV-19/Switzerland/BS-UHB-42195727/2020\|EPI_ISL_5 | ···················· | ···················· | ···················· | ···················· | ···················· | ···················· |
| G | 8901 hCoV-19/Switzerland/BS-UHB-42195791/2020\|EPI_ISL_5 | ···················· | ···················· | ···················· | ···················· | ···················· | ···················· |
| G | 8902 hCoV-19/Switzerland/BS-UHB-42195828/2020\|EPI_ISL_5 | ···················· | ···················· | ···················· | ···················· | ···················· | ···················· |
| G | 8903 hCoV-19/Switzerland/BL-UHB-42195888/2020\|EPI_ISL_5 | ···················· | ···················· | ···················· | ···················· | ···················· | ···················· |
| G | 8904 hCoV-19/Switzerland/BS-UHB-42195947/2020\|EPI_ISL_5 | ···················· | ···················· | ···················· | ···················· | ···················· | ···················· |
| G | 8905 hCoV-19/Switzerland/BS-UHB-42195951/2020\|EPI_ISL_5 | ···················· | ···················· | ···················· | ···················· | ···················· | ···················· |
| G | 8906 hCoV-19/Switzerland/BL-UHB-42196011/2020\|EPI_ISL_5 | ···················· | ·A·················· | ···················· | ···················· | ···················· | ···················· |
| G | 8907 hCoV-19/Switzerland/BS-UHB-42196028/2020\|EPI_ISL_5 | ···················· | ···················· | ···················· | ···················· | ···················· | ···················· |
| G | 8908 hCoV-19/Switzerland/BS-UHB-42196052/2020\|EPI_ISL_5 | ···················· | ···················· | ···················· | ···················· | ···················· | ···················· |
| G | 8909 hCoV-19/Switzerland/BS-UHB-42196271/2020\|EPI_ISL_5 | ···················· | ···················· | ···················· | ···················· | ···················· | ···················· |
| G | 8910 hCoV-19/Switzerland/BL-UHB-42196956/2020\|EPI_ISL_5 | ···················· | ···················· | ···················· | ···················· | ···················· | ···················· |
| G | 8911 hCoV-19/Switzerland/BS-UHB-42202260/2020\|EPI_ISL_5 | ···················· | ···················· | ···················· | ···················· | ···················· | ···················· |
| G | 8912 hCoV-19/Switzerland/BS-UHB-42202280/2020\|EPI_ISL_5 | ···················· | ···················· | ···················· | ···················· | ···················· | ···················· |
| G | 8913 hCoV-19/Switzerland/BS-UHB-42202292/2020\|EPI_ISL_5 | ···················· | ···················· | ···················· | ···················· | ···················· | ···················· |
| G | 8915 hCoV-19/Switzerland/BS-UHB-42202970/2020\|EPI_ISL_5 | ···················· | ···················· | ···················· | ···················· | ···················· | ···················· |
| G | 8916 hCoV-19/Switzerland/BS-UHB-42203020/2020\|EPI_ISL_5 | ···················· | ···················· | ···················· | ···················· | ···················· | ···················· |
| G | 8917 hCoV-19/Switzerland/BS-UHB-42203098/2020\|EPI_ISL_5 | ···················· | ···················· | ···················· | ···················· | ···················· | ···················· |
| G | 8918 hCoV-19/Switzerland/BS-UHB-42203106/2020\|EPI_ISL_5 | ···················· | ···················· | ···················· | ···················· | ···················· | ···················· |
| G | 8919 hCoV-19/Switzerland/BS-UHB-42203155/2020\|EPI_ISL_5 | ···················· | ···················· | ···················· | ···················· | ···················· | ···················· |
| G | 8920 hCoV-19/Switzerland/BS-UHB-42203196/2020\|EPI_ISL_5 | ···················· | ···················· | ···················· | ···················· | ···················· | ···················· |
| G | 8921 hCoV-19/Switzerland/BS-UHB-42203282/2020\|EPI_ISL_5 | ···················· | ···················· | ···················· | ···················· | ···················· | ···················· |
| G | 8922 hCoV-19/Switzerland/BS-UHB-42203345/2020\|EPI_ISL_5 | ···················· | ···················· | ···················· | ···················· | ··N················· | ···················· |
| G | 8923 hCoV-19/Switzerland/BS-UHB-42203355/2020\|EPI_ISL_5 | ···················· | ···················· | ···················· | ···················· | ···················· | ···················· |
| G | 8924 hCoV-19/Switzerland/BS-UHB-42203366/2020\|EPI_ISL_5 | ···················· | ···················· | ···················· | ···················· | ···················· | ···················· |
| G | 8925 hCoV-19/Switzerland/BL-UHB-42203375/2020\|EPI_ISL_5 | ···················· | ···················· | ···················· | ···················· | ···················· | ···················· |
| G | 8926 hCoV-19/Switzerland/BS-UHB-42203381/2020\|EPI_ISL_5 | ···················· | ···················· | ···················· | ···················· | ···················· | ···················· |
| G | 8928 hCoV-19/Switzerland/BS-UHB-42203579/2020\|EPI_ISL_5 | ···················· | ···················· | ···················· | ···················· | ···················· | ···················· |

| | | | | | | | | |
|---|---|---|---|---|---|---|---|---|
| G | 8929 hCoV-19/Switzerland/BS-UHB-42203802/2020\|EPI_ISL_5 | .................... | .................... | .................... | .................... | .................... | .................... | |
| G | 8930 hCoV-19/England/CAMB-1B5C41/2020\|EPI_ISL_529660\|20 | .................... | .................... | .................... | .................... | .................... | .................... | |
| G | 8931 hCoV-19/England/HECH-259A47D/2020\|EPI_ISL_529661\|2 | .................... | .................... | .................... | .................... | .................... | .................... | |
| G | 8932 hCoV-19/England/HECH-259A4AA/2020\|EPI_ISL_529662\|2 | .................... | .................... | .................... | .................... | .................... | .................... | |
| G | 8933 hCoV-19/England/HECH-259A4F5/2020\|EPI_ISL_529663\|2 | .................... | .................... | .................... | .................... | .................... | .................... | |
| G | 8934 hCoV-19/England/HECH-259A501/2020\|EPI_ISL_529664\|2 | .................... | .................... | .................... | .................... | .................... | .................... | |
| G | 8935 hCoV-19/England/HECH-259A53E/2020\|EPI_ISL_529665\|2 | .................... | .................... | .................... | .................... | .................... | .................... | |
| G | 8936 hCoV-19/England/HECH-259A54D/2020\|EPI_ISL_529666\|2 | .................... | .................... | .................... | .................... | .................... | .................... | |
| G | 8937 hCoV-19/England/NOTT-112FA3/2020\|EPI_ISL_529667\|20 | .................... | .................... | .................... | .................... | .................... | .................... | |
| G | 8938 hCoV-19/England/PORT-2D2B6E/2020\|EPI_ISL_529668\|20 | .................... | .................... | .................... | .................... | .................... | .................... | |
| G | 8939 hCoV-19/England/PORT-2D3347/2020\|EPI_ISL_529669\|20 | .................... | .................... | .................... | .................... | .................... | .................... | |
| G | 8940 hCoV-19/England/PORT-2D3383/2020\|EPI_ISL_529670\|20 | .................... | .A.................. | .................... | .................... | .................... | .................... | |
| G | 8941 hCoV-19/England/PORT-2D33DE/2020\|EPI_ISL_529671\|20 | .................... | .................... | .................... | .................... | .................... | .................... | |
| G | 8942 hCoV-19/England/PORT-2D38FD/2020\|EPI_ISL_529672\|20 | .................... | .................... | .................... | .................... | .................... | .................... | |
| G | 8943 hCoV-19/England/PORT-2D3A42/2020\|EPI_ISL_529673\|20 | .................... | .................... | .................... | .................... | .................... | .................... | |
| G | 8944 hCoV-19/England/PORT-2D40F4/2020\|EPI_ISL_529674\|20 | .................... | .................... | .................... | .................... | .................... | .................... | |
| G | 8945 hCoV-19/England/SHEF-C1092/2020\|EPI_ISL_529675\|202 | .................... | .................... | .................... | .................... | .................... | .................... | |
| G | 8946 hCoV-19/England/SHEF-C13B7/2020\|EPI_ISL_529676\|202 | .................... | .................... | .................... | .................... | .................... | .................... | |
| G | 8947 hCoV-19/England/SHEF-C1609/2020\|EPI_ISL_529677\|202 | .................... | .................... | .................... | .................... | .................... | .................... | |
| G | 8948 hCoV-19/England/SHEF-C4CB9/2020\|EPI_ISL_529678\|202 | .................... | .................... | .................... | .................... | .................... | .................... | |
| G | 8949 hCoV-19/England/SHEF-C665F/2020\|EPI_ISL_529679\|202 | .................... | .................... | .................... | .................... | .................... | .................... | |
| G | 8950 hCoV-19/England/SHEF-C666E/2020\|EPI_ISL_529680\|202 | .................... | .................... | .................... | .................... | .................... | .................... | |
| G | 8951 hCoV-19/England/SHEF-C6EB1/2020\|EPI_ISL_529681\|202 | .................... | .................... | .................... | .................... | .................... | .................... | |
| G | 8952 hCoV-19/England/SHEF-C6F45/2020\|EPI_ISL_529682\|202 | .................... | .................... | .................... | .................... | .................... | .................... | |
| G | 8953 hCoV-19/England/SHEF-CE1A1/2020\|EPI_ISL_529683\|202 | .................... | .................... | .................... | .................... | .................... | .................... | |
| G | 8954 hCoV-19/Scotland/CVR3970/2020\|EPI_ISL_529684\|2020- | .................... | .A.................. | .................... | .................... | .................... | .................... | |
| G | 8955 hCoV-19/Scotland/CVR3971/2020\|EPI_ISL_529685\|2020- | .................... | .................... | .................... | .................... | .................... | .................... | |
| G | 8956 hCoV-19/Scotland/CVR3974/2020\|EPI_ISL_529686\|2020- | .................... | .................... | .................... | .................... | .................... | .................... | |
| G | 8957 hCoV-19/Scotland/CVR3976/2020\|EPI_ISL_529687\|2020- | .................... | .................... | .................... | .................... | .................... | ...N..NN............ | 1 |
| G | 8958 hCoV-19/Scotland/CVR3977/2020\|EPI_ISL_529688\|2020- | .................... | .................... | .................... | .................... | .................... | .................... | |
| G | 8959 hCoV-19/Scotland/EDB3599/2020\|EPI_ISL_529689\|2020- | .................... | .................... | .................... | .................... | .................... | .................... | |
| G | 8960 hCoV-19/Scotland/EDB5456/2020\|EPI_ISL_529690\|2020- | .................... | .................... | .................... | .................... | .................... | .................... | |
| G | 8961 hCoV-19/England/LIVE-AA81F/2020\|EPI_ISL_517508\|202 | .................... | .................... | .................... | .................... | .................... | .................... | |
| G | 8962 hCoV-19/England/LIVE-AA82E/2020\|EPI_ISL_517509\|202 | .................... | .................... | .................... | .................... | .................... | .................... | |
| G | 8963 hCoV-19/England/PORT-2D2C3E/2020\|EPI_ISL_517517\|20 | .................... | .................... | .................... | .................... | .................... | .................... | |
| G | 8964 hCoV-19/England/PORT-2D2C6B/2020\|EPI_ISL_517519\|20 | .................... | .................... | .................... | .................... | .................... | .................... | |
| G | 8965 hCoV-19/England/PORT-2D330B/2020\|EPI_ISL_517522\|20 | .................... | .................... | .................... | .................... | .................... | .................... | |
| G | 8966 hCoV-19/England/PORT-2D33A1/2020\|EPI_ISL_517526\|20 | .................... | .................... | .................... | .................... | .................... | .................... | |
| G | 8967 hCoV-19/England/PORT-2D363F/2020\|EPI_ISL_517529\|20 | .................... | .................... | .................... | .................... | .................... | .................... | |
| G | 8968 hCoV-19/Peru/ICA-INS-055/2020\|EPI_ISL_517531\|2020- | .................... | .................... | .................... | .................... | .................... | .................... | |
| G | 8969 hCoV-19/England/PORT-2D3699/2020\|EPI_ISL_517532\|20 | .................... | .................... | .................... | .................... | .................... | .................... | |
| G | 8970 hCoV-19/England/PORT-2D36C6/2020\|EPI_ISL_517535\|20 | .................... | .................... | .................... | .................... | .................... | .................... | |
| G | 8971 hCoV-19/England/PORT-2D36D5/2020\|EPI_ISL_517536\|20 | .................... | .................... | .................... | .................... | .................... | .................... | |
| G | 8972 hCoV-19/England/PORT-2D373C/2020\|EPI_ISL_517537\|20 | .................... | .................... | .................... | .................... | .................... | .................... | |
| G | 8973 hCoV-19/England/PORT-2D3778/2020\|EPI_ISL_517540\|20 | .................... | .................... | .................... | .................... | .................... | .................... | |
| G | 8974 hCoV-19/England/PORT-2D3C1F/2020\|EPI_ISL_517542\|20 | .................... | .................... | .................... | .................... | .................... | .................... | |
| G | 8975 hCoV-19/England/SHEF-C0D05/2020\|EPI_ISL_517543\|202 | .................... | .................... | .................... | .................... | .................... | .................... | |
| G | 8976 hCoV-19/England/SHEF-C0D41/2020\|EPI_ISL_517544\|202 | .................... | .................... | .................... | .................... | .................... | .................... | |
| G | 8977 hCoV-19/England/SHEF-C33B5/2020\|EPI_ISL_517546\|202 | .................... | .................... | .................... | .................... | .................... | .................... | |
| G | 8978 hCoV-19/England/SHEF-C3AED/2020\|EPI_ISL_517548\|202 | .................... | .................... | .................... | .................... | .................... | .................... | |
| G | 8979 hCoV-19/England/SHEF-C527A/2020\|EPI_ISL_517550\|202 | .................... | .................... | .................... | .................... | .................... | .................... | |
| G | 8980 hCoV-19/England/SHEF-C585A/2020\|EPI_ISL_517552\|202 | .................... | .................... | .................... | .................... | .................... | .................... | |
| G | 8981 hCoV-19/England/SHEF-C71F3/2020\|EPI_ISL_517556\|202 | .................... | .................... | .................... | .................... | .................... | .................... | |
| G | 8982 hCoV-19/Oman/RESP-20-B-16239/2020\|EPI_ISL_518839\|2 | .................... | .................... | .................... | .................... | .................... | .................... | |
| G | 8983 hCoV-19/Oman/RESP-20-B-16300/2020\|EPI_ISL_518842\|2 | .................... | .................... | .................... | .................... | .................... | .................... | |
| G | 8984 hCoV-19/Oman/RESP-20-B-2847/2020\|EPI_ISL_518843\|20 | .................... | .................... | .................... | .................... | .................... | .................... | |
| G | 8985 hCoV-19/Oman/RESP-20-B-2876/2020\|EPI_ISL_518844\|20 | .................... | .................... | .................... | .................... | .................... | .................... | |
| G | 8986 hCoV-19/Oman/RESP-20-B-2911/2020\|EPI_ISL_518845\|20 | .................... | .................... | .................... | .................... | .................... | .................... | |
| G | 8987 hCoV-19/Oman/RESP-20-B-2932/2020\|EPI_ISL_518847\|20 | .................... | .................... | .................... | .................... | .................... | .................... | |
| G | 8988 hCoV-19/Oman/RESP-20-B-2944/2020\|EPI_ISL_518849\|20 | .................... | .................... | .................... | .................... | .................... | .................... | |
| G | 8989 hCoV-19/Oman/RESP-20-B-2964/2020\|EPI_ISL_518850\|20 | .................... | .................... | .................... | .................... | .................... | .................... | |
| G | 8990 hCoV-19/Oman/RESP-20-B-3561/2020\|EPI_ISL_518851\|20 | .................... | .................... | .................... | .................... | .................... | .................... | |
| G | 8991 hCoV-19/Oman/RESP-20-B-3562/2020\|EPI_ISL_518852\|20 | .................... | .................... | .................... | .................... | .................... | .................... | |
| G | 8992 hCoV-19/Oman/RESP-20-B-3563/2020\|EPI_ISL_518853\|20 | .................... | .................... | .................... | .................... | .................... | .................... | |
| G | 8993 hCoV-19/Oman/RESP-20-B-3564/2020\|EPI_ISL_518854\|20 | .................... | .................... | .................... | .................... | .................... | .................... | |
| G | 8994 hCoV-19/USA/MN-MDH-1461/2020\|EPI_ISL_518855\|2020-0 | .................... | .................... | .................... | .................... | .................... | .................... | |
| G | 8995 hCoV-19/USA/MN-MDH-1462/2020\|EPI_ISL_518856\|2020-0 | .................... | .................... | .................... | .................... | .................... | .................... | |
| G | 8996 hCoV-19/USA/MN-MDH-1463/2020\|EPI_ISL_518857\|2020-0 | .................... | .................... | .................... | .................... | .................... | .................... | |
| G | 8997 hCoV-19/USA/MN-MDH-1464/2020\|EPI_ISL_518858\|2020-0 | .................... | .................... | .................... | .................... | .................... | .................... | |
| G | 8998 hCoV-19/USA/MN-MDH-1465/2020\|EPI_ISL_518859\|2020-0 | .................... | .................... | .................... | .................... | .................... | .................... | |
| G | 8999 hCoV-19/USA/MN-MDH-1466/2020\|EPI_ISL_518860\|2020-0 | .................... | .................... | .................... | .................... | .................... | .................... | |
| G | 9000 hCoV-19/USA/MN-MDH-1467/2020\|EPI_ISL_518861\|2020-0 | .................... | .................... | .................... | .................... | .................... | .................... | |
| G | 9001 hCoV-19/USA/MN-MDH-1468/2020\|EPI_ISL_518862\|2020-0 | .................... | .................... | .................... | .................... | .................... | .................... | |
| G | 9002 hCoV-19/USA/MN-MDH-1469/2020\|EPI_ISL_518863\|2020-0 | .................... | .................... | .................... | .................... | .................... | .................... | |
| G | 9003 hCoV-19/USA/MN-MDH-1470/2020\|EPI_ISL_518864\|2020-0 | .................... | .................... | .................... | .................... | .................... | .................... | |
| G | 9004 hCoV-19/USA/MN-MDH-1471/2020\|EPI_ISL_518865\|2020-0 | .................... | .................... | .................... | .................... | .................... | .................... | |
| G | 9005 hCoV-19/USA/MN-MDH-1472/2020\|EPI_ISL_518866\|2020-0 | .................... | .................... | .................... | .................... | .................... | .................... | |
| G | 9006 hCoV-19/USA/MN-MDH-1473/2020\|EPI_ISL_518867\|2020-0 | .................... | .................... | .................... | .................... | .................... | .................... | |
| G | 9007 hCoV-19/USA/MN-MDH-1474/2020\|EPI_ISL_518868\|2020-0 | .................... | .................... | .................... | .................... | .................... | .................... | |
| G | 9008 hCoV-19/USA/MN-MDH-1475/2020\|EPI_ISL_518869\|2020-0 | .................... | .................... | .................... | .................... | .................... | .................... | |

G 9009 hCoV-19/USA/MN-MDH-1476/2020|EPI_ISL_518870|2020-0 ··················· ··················· ··················· ··················· ··················· ···················
G 9010 hCoV-19/USA/MN-MDH-1477/2020|EPI_ISL_518871|2020-0 ··················· ··················· ··················· ··················· ··················· ···················
G 9011 hCoV-19/USA/MN-MDH-1478/2020|EPI_ISL_518872|2020-0 ··················· ··················· ··················· ··················· ··················· ···················
G 9012 hCoV-19/USA/MN-MDH-1479/2020|EPI_ISL_518873|2020-0 ··················· ··················· ··················· ··················· ··················· ···················
G 9013 hCoV-19/USA/MN-MDH-1480/2020|EPI_ISL_518874|2020-0 ··················· ··················· ··················· ··················· ··················· ···················
G 9014 hCoV-19/USA/MN-MDH-1481/2020|EPI_ISL_518875|2020-0 ··················· ··················· ··················· ··················· ··················· ···················
G 9015 hCoV-19/USA/MN-MDH-1482/2020|EPI_ISL_518876|2020-0 ··················· ··················· ··················· ··················· ··················· ···················
G 9016 hCoV-19/USA/MN-MDH-1483/2020|EPI_ISL_518877|2020-0 ··················· ··················· ··················· ··················· ··················· ···················
G 9017 hCoV-19/USA/MN-MDH-1484/2020|EPI_ISL_518878|2020-0 ··················· ··················· ··················· ··················· ··················· ···················
G 9018 hCoV-19/USA/MN-MDH-1485/2020|EPI_ISL_518879|2020-0 ··················· ··················· ··················· ··················· ··················· ···················
G 9019 hCoV-19/USA/MN-MDH-1487/2020|EPI_ISL_518880|2020-0 ··················· ··················· ··················· ··················· ··················· ···················
G 9020 hCoV-19/USA/MN-MDH-1488/2020|EPI_ISL_518881|2020-0 ··················· ··················· ··················· ··················· ··················· ···················
G 9021 hCoV-19/USA/MN-MDH-1489/2020|EPI_ISL_518882|2020-0 ··················· ··················· ··················· ··················· ··················· ···················
G 9022 hCoV-19/USA/MN-MDH-1490/2020|EPI_ISL_518883|2020-0 ··················· ··················· ··················· ··················· ··················· ···················
G 9023 hCoV-19/USA/MN-MDH-1491/2020|EPI_ISL_518884|2020-0 ··················· ··················· ··················· ··················· ··················· ···················
G 9024 hCoV-19/USA/MN-MDH-1492/2020|EPI_ISL_518885|2020-0 ··················· ··················· ··················· ··················· ··················· ···················
G 9025 hCoV-19/USA/MN-MDH-1493/2020|EPI_ISL_518886|2020-0 ··················· ··················· ··················· ··················· ··················· ···················
G 9026 hCoV-19/USA/MN-MDH-1494/2020|EPI_ISL_518887|2020-0 ··················· ··················· ··················· ··················· ··················· ···················
G 9027 hCoV-19/USA/MN-MDH-1495/2020|EPI_ISL_518888|2020-0 ··················· ··················· ··················· ··················· ··················· ···················
G 9028 hCoV-19/USA/MN-MDH-1496/2020|EPI_ISL_518889|2020-0 ··················· ··················· ··················· ··················· ··················· ···················
G 9030 hCoV-19/Australia/VIC2279/2020|EPI_ISL_522001|2020 ··················· ··················· ··················· ··················· ··················· ···················
G 9031 hCoV-19/Australia/VIC2280/2020|EPI_ISL_522002|2020 ··················· ··················· ··················· ··················· ··················· ···················
G 9032 hCoV-19/Australia/VIC2281/2020|EPI_ISL_522003|2020 ··················· ··················· ··················· ··················· ··················· ···················
G 9033 hCoV-19/Australia/VIC2282/2020|EPI_ISL_522004|2020 ··················· ··················· ··················· ··················· ··················· ···················
G 9034 hCoV-19/Australia/VIC2283/2020|EPI_ISL_522005|2020 ··················· ··················· ··················· ··················· ··················· ···················
G 9035 hCoV-19/Australia/VIC2284/2020|EPI_ISL_522006|2020 ··················· ··················· ··················· ··················· ··················· ···················
G 9036 hCoV-19/Australia/VIC2285/2020|EPI_ISL_522007|2020 ··················· ··················· ··················· ··················· ··················· ···················
G 9037 hCoV-19/Australia/VIC2286/2020|EPI_ISL_522008|2020 ··················· ··················· ··················· ··················· ··················· ···················
G 9038 hCoV-19/Australia/VIC2287/2020|EPI_ISL_522009|2020 ··················· ··················· ··················· ··················· ··················· ···················
G 9039 hCoV-19/Australia/VIC2288/2020|EPI_ISL_522010|2020 ··················· ··················· ··················· ··················· ··················· ···················
G 9040 hCoV-19/Australia/VIC2289/2020|EPI_ISL_522011|2020 ··················· ··················· ··················· ··················· ··················· ···················
G 9042 hCoV-19/Australia/VIC2291/2020|EPI_ISL_522013|2020 ··················· ··················· ··················· ··················· ··················· ···················
G 9043 hCoV-19/Australia/VIC2292/2020|EPI_ISL_522014|2020 ··················· ··················· ··················· ··················· ··················· ···················
G 9044 hCoV-19/Australia/VIC2294/2020|EPI_ISL_522015|2020 ··················· ··················· ··················· ··················· N·················· ···················
G 9045 hCoV-19/Australia/VIC2295/2020|EPI_ISL_522016|2020 ··················· ··················· ··················· ··················· ··················· ···················
G 9046 hCoV-19/Australia/VIC2296/2020|EPI_ISL_522017|2020 ··················· ··················· ··················· ··················· ··················· ···················
G 9047 hCoV-19/Australia/VIC2297/2020|EPI_ISL_522018|2020 ··················· ··················· ··················· ··················· NNN················ ··················· 1
G 9048 hCoV-19/Australia/VIC2298/2020|EPI_ISL_522019|2020 ··················· ··················· ··················· ··················· ··················· ···················
G 9050 hCoV-19/Australia/VIC2300/2020|EPI_ISL_522021|2020 ··················· ··················· ··················· ··················· ··················· ···················
G 9052 hCoV-19/Australia/VIC2302/2020|EPI_ISL_522023|2020 ··················· ··················· ··················· ··················· ··················· ···················
G 9054 hCoV-19/Australia/VIC2304/2020|EPI_ISL_522025|2020 ··················· ··················· ··················· ··················· ··················· ···················
G 9055 hCoV-19/Australia/VIC2305/2020|EPI_ISL_522026|2020 ··················· ··················· ··················· ··················· ··················· ···················
G 9056 hCoV-19/Australia/VIC2306/2020|EPI_ISL_522027|2020 ··················· ··················· ··················· ··················· ··················· ···················
G 9058 hCoV-19/Australia/VIC2308/2020|EPI_ISL_522029|2020 ··················· ··················· ··················· ··················· ··················· ···················
G 9059 hCoV-19/Australia/VIC2309/2020|EPI_ISL_522030|2020 ··················· ··················· ··················· ··················· ··················· ···················
G 9064 hCoV-19/Australia/VIC2314/2020|EPI_ISL_522035|2020 ··················· ··················· ··················· ··················· ··NN··············· ···················
G 9065 hCoV-19/Australia/VIC2315/2020|EPI_ISL_522036|2020 ··················· ··················· ··················· ··················· NNN················ ··················· 1
G 9066 hCoV-19/Australia/VIC2316/2020|EPI_ISL_522037|2020 ··················· ··················· ··················· ··················· ··················· ···················
G 9067 hCoV-19/Australia/VIC2317/2020|EPI_ISL_522038|2020 ··················· ··················· ··················· ··················· ·NN················ ···················
G 9070 hCoV-19/Australia/VIC2320/2020|EPI_ISL_522041|2020 ··················· ··················· ··················· ··················· ··················· ···················
G 9071 hCoV-19/Australia/VIC2321/2020|EPI_ISL_522042|2020 ··················· ··················· ··················· ··················· ··················· ···················
G 9072 hCoV-19/Australia/VIC2322/2020|EPI_ISL_522043|2020 ··················· ··················· ··················· ··················· ··················· ···················
G 9073 hCoV-19/Australia/VIC2323/2020|EPI_ISL_522044|2020 ··················· ··················· ··················· ··················· ··················· ···················
G 9074 hCoV-19/Australia/VIC2324/2020|EPI_ISL_522045|2020 ··················· ··················· ··················· ··················· ··················· ···················
G 9075 hCoV-19/Australia/VIC2325/2020|EPI_ISL_522046|2020 ··················· ··················· ··················· ··················· ··················· ···················
G 9076 hCoV-19/Australia/VIC2326/2020|EPI_ISL_522047|2020 ··················· ··················· ··················· ··················· ··················· ···················
G 9077 hCoV-19/Australia/VIC2327/2020|EPI_ISL_522048|2020 ··················· ··················· ··················· ··················· ··················· ···················
G 9079 hCoV-19/Australia/VIC2333/2020|EPI_ISL_522054|2020 ··················· ··················· ··················· ··················· ··················· ···················
G 9080 hCoV-19/Australia/VIC2334/2020|EPI_ISL_522055|2020 ··················· ··················· ··················· ··················· ··················· ·NNN··············· 1
G 9081 hCoV-19/Australia/VIC2335/2020|EPI_ISL_522056|2020 ··················· ··················· ··················· ··················· ··················· ···················
G 9082 hCoV-19/Australia/VIC2336/2020|EPI_ISL_522057|2020 ··················· ··················· ··················· ··················· ··················· ···················
G 9083 hCoV-19/Australia/VIC2337/2020|EPI_ISL_522058|2020 ··················· ··················· ··················· ··················· ··················· ···················
G 9084 hCoV-19/Australia/VIC2338/2020|EPI_ISL_522059|2020 ··················· ··················· ··················· ··················· ··················· ···················
G 9085 hCoV-19/Netherlands/NB-EMC-193/2020|EPI_ISL_523316 ··················· ··················· ··················· ··················· ··················· ···················
G 9086 hCoV-19/Netherlands/NB-EMC-194/2020|EPI_ISL_523317 ··················· ··················· ··················· ··················· ··················· ···················
G 9087 hCoV-19/Netherlands/NB-EMC-195/2020|EPI_ISL_523318 ··················· ··················· ··················· ··················· ··················· ···················
G 9088 hCoV-19/Netherlands/NB-EMC-196/2020|EPI_ISL_523319 ··················· ··················· ··················· ··················· ··················· ···················
G 9089 hCoV-19/Netherlands/NB-EMC-197/2020|EPI_ISL_523320 ··················· ··················· ··················· ··················· ··················· ···················
G 9090 hCoV-19/Netherlands/NB-EMC-198/2020|EPI_ISL_523321 ··················· ··················· ··················· ··················· ··················· ···················
G 9091 hCoV-19/Netherlands/NB-EMC-199/2020|EPI_ISL_523322 ··················· ··················· ··················· ··················· ··················· ···················
G 9092 hCoV-19/Netherlands/NB-EMC-200/2020|EPI_ISL_523323 ··················· ··················· ··················· ··················· ··················· ···················
G 9093 hCoV-19/Netherlands/NB-EMC-201/2020|EPI_ISL_523324 ··················· ··················· ··················· ··················· ··················· ···················
G 9094 hCoV-19/Netherlands/NB-EMC-202/2020|EPI_ISL_523325 ··················· ··················· ··················· ··················· ··················· ···················
G 9095 hCoV-19/Netherlands/NB-EMC-203/2020|EPI_ISL_523326 ··················· ··················· ··················· ··················· ··················· ···················
G 9096 hCoV-19/Netherlands/NB-EMC-204/2020|EPI_ISL_523327 ··················· ··················· ··················· ··················· ··················· ···················
G 9097 hCoV-19/Netherlands/NB-EMC-205/2020|EPI_ISL_523328 ··················· ··················· ··················· ··················· ··················· ···················
G 9098 hCoV-19/Netherlands/NB-EMC-206/2020|EPI_ISL_523329 ··················· ··················· ··················· ··················· ··················· ···················
G 9099 hCoV-19/Netherlands/NB-EMC-207/2020|EPI_ISL_523330 ··················· ··················· ··················· ··················· ··················· ···················
G 9100 hCoV-19/Netherlands/NB-EMC-208/2020|EPI_ISL_523331 ··················· ··················· ··················· ··················· ··················· ···················
G 9101 hCoV-19/Netherlands/NB-EMC-209/2020|EPI_ISL_523332 ··················· ··················· ··················· ··················· ··················· ···················

| | | | | | | | | |
|---|---|---|---|---|---|---|---|---|
| G | 9102 | hCoV-19/Netherlands/NB-EMC-210/2020\|EPI_ISL_523333 | ···················· | ···················· | ···················· | ···················· | ···················· | ···················· |
| G | 9103 | hCoV-19/Netherlands/NB-EMC-211/2020\|EPI_ISL_523334 | ···················· | ···················· | ···················· | ···················· | ···················· | ···················· |
| G | 9104 | hCoV-19/Netherlands/NB-EMC-212/2020\|EPI_ISL_523335 | ···················· | ·A·················· | ···················· | ···················· | ···················· | ···A················ |
| G | 9105 | hCoV-19/Netherlands/NB-EMC-213/2020\|EPI_ISL_523336 | ···················· | ·A·················· | ···················· | ···················· | ···················· | ···A················ |
| G | 9106 | hCoV-19/Netherlands/NB-EMC-214/2020\|EPI_ISL_523337 | ···················· | ···················· | ···················· | ···················· | ···················· | ···················· |
| G | 9107 | hCoV-19/Netherlands/NB-EMC-217/2020\|EPI_ISL_523340 | ···················· | ·A·················· | ···················· | ···················· | ···················· | ···A················ |
| G | 9108 | hCoV-19/Netherlands/NB-EMC-218/2020\|EPI_ISL_523341 | ···················· | ·A·················· | ···················· | ···················· | ···················· | ···A················ |
| G | 9109 | hCoV-19/Netherlands/NB-EMC-219/2020\|EPI_ISL_523342 | ···················· | ·A·················· | ···················· | ···················· | ···················· | ···A················ |
| G | 9110 | hCoV-19/Netherlands/NB-EMC-220/2020\|EPI_ISL_523343 | ···················· | ·A·················· | ···················· | ···················· | ···················· | ···A················ |
| G | 9111 | hCoV-19/Netherlands/NB-EMC-222/2020\|EPI_ISL_523345 | ···················· | ···················· | ···················· | ···················· | ···················· | ···················· |
| G | 9112 | hCoV-19/Netherlands/NB-EMC-224/2020\|EPI_ISL_523347 | ···················· | ···················· | ···················· | ···················· | ···················· | ···················· |
| G | 9113 | hCoV-19/Netherlands/NB-EMC-225/2020\|EPI_ISL_523348 | ···················· | ···················· | ···················· | ···················· | ···················· | ···················· |
| G | 9114 | hCoV-19/Netherlands/NB-EMC-226/2020\|EPI_ISL_523349 | ···················· | ···················· | ···················· | ···················· | ···················· | ···················· |
| G | 9115 | hCoV-19/Netherlands/NB-EMC-227/2020\|EPI_ISL_523350 | ···················· | ···················· | ···················· | ···················· | ···················· | ···················· |
| G | 9116 | hCoV-19/Netherlands/NB-EMC-228/2020\|EPI_ISL_523351 | ···················· | ···················· | ···················· | ···················· | ···················· | ···················· |
| G | 9117 | hCoV-19/Netherlands/NB-EMC-229/2020\|EPI_ISL_523352 | ···················· | ···················· | ···················· | ···················· | ···················· | ···················· |
| G | 9118 | hCoV-19/Netherlands/NB-EMC-230/2020\|EPI_ISL_523353 | ···················· | ···················· | ···················· | ···················· | ···················· | ···················· |
| G | 9119 | hCoV-19/Netherlands/NB-EMC-231/2020\|EPI_ISL_523354 | ···················· | ···················· | ···················· | ···················· | ···················· | ···················· |
| G | 9120 | hCoV-19/Netherlands/NB-EMC-232/2020\|EPI_ISL_523355 | ···················· | ···················· | ···················· | ···················· | ···················· | ···················· |
| G | 9121 | hCoV-19/Netherlands/NB-EMC-233/2020\|EPI_ISL_523356 | ···················· | ···················· | ···················· | ···················· | ···················· | ···················· |
| G | 9122 | hCoV-19/Netherlands/NB-EMC-234/2020\|EPI_ISL_523357 | ···················· | ···················· | ···················· | ···················· | ···················· | ···················· |
| G | 9123 | hCoV-19/Netherlands/NB-EMC-235/2020\|EPI_ISL_523358 | ···················· | ···················· | ···················· | ···················· | ···················· | ···················· |
| G | 9124 | hCoV-19/Netherlands/NB-EMC-236/2020\|EPI_ISL_523359 | ···················· | ···················· | ···················· | ···················· | ···················· | ···················· |
| G | 9125 | hCoV-19/Netherlands/NB-EMC-238/2020\|EPI_ISL_523361 | ···················· | ···················· | ···················· | ···················· | ···················· | ···················· |
| G | 9126 | hCoV-19/Netherlands/NB-EMC-241/2020\|EPI_ISL_523364 | ···················· | ···················· | ···················· | ···················· | ···················· | ···················· |
| G | 9127 | hCoV-19/Netherlands/NB-EMC-243/2020\|EPI_ISL_523366 | ···················· | ···················· | ···················· | ···················· | ···················· | ···················· |
| G | 9128 | hCoV-19/Netherlands/NB-EMC-244/2020\|EPI_ISL_523367 | ···················· | ···················· | ···················· | ···················· | ···················· | ···················· |
| G | 9129 | hCoV-19/Netherlands/NB-EMC-245/2020\|EPI_ISL_523368 | ···················· | ···················· | ···················· | ···················· | ···················· | ···················· |
| G | 9130 | hCoV-19/Netherlands/NB-EMC-246/2020\|EPI_ISL_523369 | ···················· | ···················· | ···················· | ···················· | ···················· | ···················· |
| G | 9131 | hCoV-19/Netherlands/NB-EMC-247/2020\|EPI_ISL_523370 | ···················· | ···················· | ···················· | ···················· | ···················· | ···················· |
| G | 9132 | hCoV-19/Netherlands/NB-EMC-248/2020\|EPI_ISL_523371 | ···················· | ···················· | ···················· | ···················· | ···················· | ···················· |
| G | 9133 | hCoV-19/Netherlands/NB-EMC-249/2020\|EPI_ISL_523372 | ···················· | ···················· | ···················· | ···················· | ···················· | ···················· |
| G | 9134 | hCoV-19/Netherlands/NB-EMC-250/2020\|EPI_ISL_523373 | ···················· | ···················· | ···················· | ···················· | ···················· | ···················· |
| G | 9135 | hCoV-19/Netherlands/NB-EMC-251/2020\|EPI_ISL_523374 | ···················· | ···················· | ···················· | ···················· | ···················· | ···················· |
| G | 9136 | hCoV-19/Netherlands/NB-EMC-252/2020\|EPI_ISL_523375 | ···················· | ···················· | ···················· | ···················· | ···················· | ···················· |
| G | 9137 | hCoV-19/Netherlands/NB-EMC-253/2020\|EPI_ISL_523376 | ···················· | ···················· | ···················· | ···················· | ···················· | ···················· |
| G | 9138 | hCoV-19/Netherlands/NB-EMC-254/2020\|EPI_ISL_523377 | ···················· | ···················· | ···················· | ···················· | ···················· | ···················· |
| G | 9139 | hCoV-19/Netherlands/NB-EMC-255/2020\|EPI_ISL_523378 | ···················· | ···················· | ···················· | ···················· | ···················· | ···················· |
| G | 9140 | hCoV-19/Netherlands/NB-EMC-262/2020\|EPI_ISL_523385 | ···················· | ···················· | ···················· | ···················· | ···················· | ···················· |
| G | 9141 | hCoV-19/Netherlands/NB-EMC-263/2020\|EPI_ISL_523386 | ···················· | ···················· | ···················· | ···················· | ···················· | ···················· |
| G | 9142 | hCoV-19/Netherlands/NB-EMC-265/2020\|EPI_ISL_523388 | ···················· | ···················· | ···················· | ···················· | ···················· | ···················· |
| G | 9143 | hCoV-19/Netherlands/NB-EMC-266/2020\|EPI_ISL_523389 | ···················· | ···················· | ···················· | ···················· | ···················· | ···················· |
| G | 9144 | hCoV-19/England/LOND-1260FDF/2020\|EPI_ISL_524647\|2 | ···················· | ···················· | ···················· | ···················· | ···················· | ···················· |
| G | 9145 | hCoV-19/England/LOND-126168E/2020\|EPI_ISL_524648\|2 | ···················· | ···················· | ···················· | ···················· | ···················· | ···················· |
| G | 9146 | hCoV-19/England/LOND-1260CBA/2020\|EPI_ISL_524649\|2 | ···················· | ···················· | ···················· | ···················· | ···················· | ···················· |
| G | 9147 | hCoV-19/England/LOND-126033D/2020\|EPI_ISL_524651\|2 | ···················· | ···················· | ···················· | ···················· | ···················· | ···················· |
| G | 9148 | hCoV-19/England/LOND-1260C8D/2020\|EPI_ISL_524652\|2 | ···················· | ···················· | ···················· | ···················· | ···················· | ···················· |
| G | 9149 | hCoV-19/England/LOND-126041C/2020\|EPI_ISL_524653\|2 | ···················· | ···················· | ···················· | ···················· | ···················· | ···················· |
| G | 9150 | hCoV-19/England/LOND-1260CAB/2020\|EPI_ISL_524654\|2 | ···················· | ···················· | ···················· | ···················· | ···················· | ···················· |
| G | 9151 | hCoV-19/England/LOND-12602B8/2020\|EPI_ISL_524656\|2 | ···················· | ···················· | ···················· | ···················· | ···················· | ···················· |
| G | 9152 | hCoV-19/England/LOND-126042B/2020\|EPI_ISL_524658\|2 | ···················· | ···················· | ···················· | ···················· | ···················· | ···················· |
| G | 9153 | hCoV-19/England/LOND-12616E8/2020\|EPI_ISL_524659\|2 | ···················· | ···················· | ···················· | ···················· | ···················· | ···················· |
| G | 9154 | hCoV-19/England/LOND-1260300/2020\|EPI_ISL_524663\|2 | ···················· | ···················· | ···················· | ···················· | ···················· | ···················· |
| G | 9155 | hCoV-19/England/LOND-1260476/2020\|EPI_ISL_524665\|2 | ···················· | ···················· | ···················· | ···················· | ···················· | ···················· |
| G | 9156 | hCoV-19/England/LOND-126078C/2020\|EPI_ISL_524668\|2 | ···················· | ···················· | ···················· | ···················· | ···················· | ···················· |
| G | 9157 | hCoV-19/England/LOND-126032E/2020\|EPI_ISL_524669\|2 | ···················· | ···················· | ···················· | ···················· | ···················· | ···················· |
| G | 9158 | hCoV-19/England/LOND-126034C/2020\|EPI_ISL_524670\|2 | ···················· | ···················· | ···················· | ···················· | ···················· | ···················· |
| G | 9159 | hCoV-19/England/LOND-126040D/2020\|EPI_ISL_524671\|2 | ···················· | ···················· | ···················· | ···················· | ···················· | ···················· |
| G | 9160 | hCoV-19/England/LOND-126035B/2020\|EPI_ISL_524673\|2 | ···················· | ···················· | ···················· | ···················· | ···················· | ···················· |
| G | 9161 | hCoV-19/England/LOND-1261642/2020\|EPI_ISL_524674\|2 | ···T················ | ···················· | ···················· | ···················· | ···················· | ···················· |
| G | 9162 | hCoV-19/England/LOND-126031F/2020\|EPI_ISL_524676\|2 | ···················· | ···················· | ···················· | ···················· | ···················· | ···················· |
| G | 9163 | hCoV-19/England/LOND-1260DA8/2020\|EPI_ISL_524680\|2 | ···················· | ···················· | ···················· | ···················· | ···················· | ···················· |
| G | 9164 | hCoV-19/England/LOND-1261026/2020\|EPI_ISL_524682\|2 | ···················· | ···················· | ···················· | ···················· | ···················· | ···················· |
| G | 9165 | hCoV-19/England/LOND-12613A5/2020\|EPI_ISL_524683\|2 | ···················· | ···················· | ···················· | ···················· | ···················· | ···················· |
| G | 9166 | hCoV-19/England/LOND-1260F48/2020\|EPI_ISL_524684\|2 | ···················· | ···················· | ···················· | ···················· | ···················· | ···················· |
| G | 9167 | hCoV-19/England/LOND-1260DB7/2020\|EPI_ISL_524686\|2 | ···················· | ···················· | ···················· | ···················· | ···················· | ···················· |
| G | 9168 | hCoV-19/England/LOND-1260FEE/2020\|EPI_ISL_524688\|2 | ···················· | ···················· | ···················· | ···················· | ···················· | ···················· |
| G | 9169 | hCoV-19/England/LOND-1260EB4/2020\|EPI_ISL_524689\|2 | ···················· | ···················· | ···················· | ···················· | ···················· | ···················· |
| G | 9170 | hCoV-19/England/LOND-12613F0/2020\|EPI_ISL_524690\|2 | ···················· | ···················· | ···················· | ···················· | ···················· | ···················· |
| G | 9171 | hCoV-19/England/LOND-12607C8/2020\|EPI_ISL_524691\|2 | ···················· | ···················· | ···················· | ···················· | ···················· | ···················· |
| G | 9172 | hCoV-19/England/LOND-126082F/2020\|EPI_ISL_524692\|2 | ···················· | ···················· | ···················· | ···················· | ···················· | ···················· |
| G | 9173 | hCoV-19/England/LOND-1261484/2020\|EPI_ISL_524693\|2 | ···················· | ···················· | ···················· | ···················· | ···················· | ···················· |
| G | 9174 | hCoV-19/England/LOND-12613B4/2020\|EPI_ISL_524694\|2 | ···················· | ···················· | ···················· | ···················· | ···················· | ···················· |
| G | 9175 | hCoV-19/England/LOND-1261590/2020\|EPI_ISL_524696\|2 | ···················· | ···················· | ···················· | ···················· | ···················· | ···················· |
| G | 9176 | hCoV-19/England/LOND-1260810/2020\|EPI_ISL_524698\|2 | ···················· | ···················· | ···················· | ···················· | ···················· | ···················· |
| G | 9177 | hCoV-19/England/LOND-126086B/2020\|EPI_ISL_524699\|2 | ···················· | ···················· | ···················· | ···················· | ···················· | ···················· |
| G | 9178 | hCoV-19/USA/OR-OHSU-1565/2020\|EPI_ISL_525979\|2020- | ···················· | ···················· | ···················· | ···················· | ···················· | ···················· |
| G | 9179 | hCoV-19/USA/OR-OHSU-1569/2020\|EPI_ISL_525980\|2020- | ···················· | ···················· | ···················· | ···················· | ···················· | ···················· |
| G | 9180 | hCoV-19/USA/OR-OHSU-1573/2020\|EPI_ISL_525981\|2020- | ···················· | ···················· | ···················· | ···················· | ···················· | ···················· |
| G | 9181 | hCoV-19/USA/OR-OHSU-1581/2020\|EPI_ISL_525982\|2020- | ···················· | ·A·················· | ···················· | ···················· | ···················· | ···················· |

G 9182 hCoV-19/USA/OR-OHSU-1583/2020|EPI_ISL_525983|2020- ···················· ···················· ····················· ···················· ···················· ····················
G 9183 hCoV-19/USA/OR-OHSU-1591/2020|EPI_ISL_525984|2020- ···················· ···················· ····················· ···················· ···················· ····················
G 9184 hCoV-19/USA/OR-OHSU-1597/2020|EPI_ISL_525985|2020- ···················· ···················· ····················· ···················· ···················· ····················
G 9185 hCoV-19/USA/OR-OHSU-1599/2020|EPI_ISL_525986|2020- ···················· ·A·················· ····················· ···················· ···················· ····················
G 9186 hCoV-19/USA/OR-OHSU-1603/2020|EPI_ISL_525987|2020- ···················· ···················· ····················· ···················· ···················· ····················
G 9187 hCoV-19/USA/OR-OHSU-1631/2020|EPI_ISL_525988|2020- ···················· ···················· ····················· ···················· ···················· ····················
G 9188 hCoV-19/USA/OR-OHSU-1635/2020|EPI_ISL_525989|2020- ···················· ···················· ····················· ···················· ···················· ····················
G 9189 hCoV-19/USA/OR-OHSU-1637/2020|EPI_ISL_525990|2020- ···················· ·A·················· ····················· ···················· ···················· ····················
G 9190 hCoV-19/USA/OR-OHSU-1643/2020|EPI_ISL_525991|2020- ···················· ·A·················· ···A················· ···················· ···················· ····················
G 9191 hCoV-19/USA/OR-OHSU-1645/2020|EPI_ISL_525992|2020- ···················· ·A·················· ····················· ···················· ···················· ····················
G 9192 hCoV-19/USA/OR-OHSU-1649/2020|EPI_ISL_525993|2020- ···················· ···················· ····················· ···················· ···················· ····················
G 9193 hCoV-19/USA/OR-OHSU-1651/2020|EPI_ISL_525994|2020- ···················· ···················· ····················· ···················· ···················· ····················
G 9194 hCoV-19/USA/OR-OHSU-1655/2020|EPI_ISL_525995|2020- ···················· ··········A········· ····················· ···················· ···················· ····················
G 9195 hCoV-19/USA/OR-OHSU-1663/2020|EPI_ISL_525996|2020- ···················· ···················· ····················· ···················· ···················· ····················
G 9196 hCoV-19/USA/OR-OHSU-1665/2020|EPI_ISL_525997|2020- ···················· ···················· ····················· ···················· ···················· ····················
G 9197 hCoV-19/USA/OR-OHSU-1673/2020|EPI_ISL_525998|2020- ···················· ·A·················· ····················· ···················· ···················· ····················
G 9198 hCoV-19/USA/OR-OHSU-1675/2020|EPI_ISL_525999|2020- ···················· ·A·················· ····················· ···················· ···················· ····················
G 9199 hCoV-19/Switzerland/BS-UHB-42196976/2020|EPI_ISL_5 ···················· ···················· ····················· ···················· ···················· ····················
G 9200 hCoV-19/Switzerland/BL-UHB-42197004/2020|EPI_ISL_5 ···················· ···················· ····················· ···················· ···················· ····················
G 9201 hCoV-19/Switzerland/BS-UHB-42197025/2020|EPI_ISL_5 ···················· ···················· ····················· ···················· ···················· ····················
G 9202 hCoV-19/Switzerland/BS-UHB-42197035/2020|EPI_ISL_5 ···················· ···················· ····················· ···················· ···················· ····················
G 9203 hCoV-19/Switzerland/BS-UHB-42197095/2020|EPI_ISL_5 ···················· ···················· ····················· ···················· ···················· ····················
G 9204 hCoV-19/Switzerland/SO-UHB-42197247/2020|EPI_ISL_5 ···················· ···················· ····················· ···················· ···················· ····················
G 9205 hCoV-19/England/ALDP-97DEA1/2020|EPI_ISL_529528|20 ···················· ···················· ····················· ···················· ···················· ····················
G 9206 hCoV-19/England/ALDP-97DEDE/2020|EPI_ISL_529529|20 ···················· ···················· ····················· ···················· ···················· ····················
G 9207 hCoV-19/England/ALDP-97F1C4/2020|EPI_ISL_529530|20 ···················· ···················· ····················· ···················· ···················· ····················
G 9208 hCoV-19/England/ALDP-97F20D/2020|EPI_ISL_529531|20 ···················· ···················· ····················· ···················· ···················· ····················
G 9209 hCoV-19/England/ALDP-97F2B2/2020|EPI_ISL_529532|20 ···················· ···················· ····················· ···················· ···················· ····················
G 9210 hCoV-19/England/ALDP-97F5B9/2020|EPI_ISL_529533|20 ···················· ···················· ····················· ···················· ···················· ····················
G 9211 hCoV-19/England/ALDP-97FB3F/2020|EPI_ISL_529534|20 ···················· ·A·················· ····················· ···················· ···················· ···A················
G 9212 hCoV-19/England/ALDP-97FC4B/2020|EPI_ISL_529535|20 ···················· ···················· ····················· ···················· ···················· ····················
G 9213 hCoV-19/England/ALDP-980236/2020|EPI_ISL_529536|20 ···················· ···················· ····················· ···················· ···················· ····················
G 9214 hCoV-19/England/ALDP-980281/2020|EPI_ISL_529537|20 ···················· ···················· ····················· ···················· ···················· ····················
G 9215 hCoV-19/England/ALDP-98056A/2020|EPI_ISL_529538|20 ···················· ···················· ····················· ···················· ···················· ····················
G 9216 hCoV-19/England/ALDP-9805B5/2020|EPI_ISL_529539|20 ···················· ···················· ····················· ···················· ···················· ····················
G 9217 hCoV-19/England/LIVE-AA79A/2020|EPI_ISL_517500|202 ···················· ···················· ····················· ···················· ···················· ····················
G 9218 hCoV-19/England/LIVE-AA7A9/2020|EPI_ISL_517501|202 ···················· ···················· ····················· ···················· ···················· ····················
G 9219 hCoV-19/England/LIVE-AA7B8/2020|EPI_ISL_517502|202 ···················· ···················· ····················· ···················· ···················· ····················
G 9220 hCoV-19/England/LIVE-AA7C7/2020|EPI_ISL_517503|202 ···················· ···················· ····················· ···················· ···················· ····················
G 9221 hCoV-19/England/LIVE-AA7D6/2020|EPI_ISL_517504|202 ···················· ···················· ····················· ···················· ···················· ····················
G 9222 hCoV-19/England/LIVE-AA7E5/2020|EPI_ISL_517505|202 ···················· ···················· ····················· ···················· ···················· ····················
G 9223 hCoV-19/England/LIVE-AA7F4/2020|EPI_ISL_517506|202 ···················· ···················· ····················· ···················· ···················· ····················
G 9224 hCoV-19/England/LIVE-AA800/2020|EPI_ISL_517507|202 ···················· ···················· ····················· ···················· ···················· ····················
G 9225 hCoV-19/England/PORT-2D2B04/2020|EPI_ISL_517512|20 ···················· ···················· ····················· ···················· ···················· ····················
G 9226 hCoV-19/England/PORT-2D2B31/2020|EPI_ISL_517513|20 ···················· ···················· ····················· ···················· ···················· ····················
G 9227 hCoV-19/England/SHEF-C9D57/2020|EPI_ISL_517560|202 ···················· ···················· ····················· ···················· ···················· ····················
G 9228 hCoV-19/England/SHEF-C9D66/2020|EPI_ISL_517561|202 ···················· ···················· ····················· ···················· ···················· ····················
G 9229 hCoV-19/England/SHEF-CF9E4/2020|EPI_ISL_517570|202 ···················· ···················· ····················· ···················· ···················· ····················
G 9230 hCoV-19/England/SHEF-CFBC0/2020|EPI_ISL_517572|202 ···················· ···················· ····················· ···················· ···················· ····················
G 9231 hCoV-19/England/SHEF-D116F/2020|EPI_ISL_517575|202 ···················· ···················· ····················· ···················· ···················· ····················
G 9232 hCoV-19/England/SHEF-D16F7/2020|EPI_ISL_517576|202 ···················· ···················· ····················· ···················· ···················· ····················
G 9233 hCoV-19/Scotland/CVR3964/2020|EPI_ISL_517580|2020- ···················· ···················· ····················· ···················· ···A················ ····················
G 9234 hCoV-19/Scotland/CVR3965/2020|EPI_ISL_517581|2020- ···················· ···················· ····················· ···················· ···················· ····················
G 9235 hCoV-19/Scotland/CVR3966/2020|EPI_ISL_517582|2020- ···················· ·A·················· ····················· ···················· ···················· ····················
G 9236 hCoV-19/Scotland/EDB6912/2020|EPI_ISL_517583|2020- ···················· ·A·················· ····················· ···················· ···················· ····················
G 9237 hCoV-19/Scotland/EDB6913/2020|EPI_ISL_517584|2020- ···················· ·A·················· ····················· ···················· ···················· ····················
G 9238 hCoV-19/Scotland/EDB6916/2020|EPI_ISL_517585|2020- ···················· ·A·················· ····················· ···················· ···················· ····················
G 9239 hCoV-19/Scotland/EDB6917/2020|EPI_ISL_517586|2020- ···················· ·A·················· ····················· ···················· ···················· ····················
G 9240 hCoV-19/Scotland/EDB6919/2020|EPI_ISL_517587|2020- ···················· ·A·················· ····················· ···················· ···················· ····················
G 9241 hCoV-19/Scotland/EDB7128/2020|EPI_ISL_517588|2020- ···················· ···················· ····················· ···················· ···················· ····················
G 9242 hCoV-19/Scotland/EDB7181/2020|EPI_ISL_517589|2020- ···················· ···················· ····················· ···················· ···················· ····················
G 9243 hCoV-19/Wales/PHWC-169B03/2020|EPI_ISL_517590|2020 ···················· ···················· ····················· ···················· ···················· ····················
G 9244 hCoV-19/Wales/PHWC-169B12/2020|EPI_ISL_517591|2020 ···················· ···················· ····················· ···················· ···················· ····················
G 9245 hCoV-19/Wales/PHWC-169B21/2020|EPI_ISL_517592|2020 ···················· ···················· ····················· ···················· ···················· ····················
G 9246 hCoV-19/Wales/PHWC-169B30/2020|EPI_ISL_517593|2020 ···················· ···················· ····················· ···················· ···················· ····················
G 9247 hCoV-19/Wales/PHWC-169B4F/2020|EPI_ISL_517594|2020 ···················· ···················· ····················· ···················· ···················· ····················
G 9248 hCoV-19/Wales/PHWC-169B5E/2020|EPI_ISL_517595|2020 ···················· ···················· ····················· ···················· ···················· ····················
G 9249 hCoV-19/Wales/PHWC-169B6D/2020|EPI_ISL_517596|2020 ···················· ···················· ····················· ···················· ···················· ····················
G 9251 hCoV-19/Wales/PHWC-169B8B/2020|EPI_ISL_517598|2020 ···················· ···················· ····················· ···················· ···················· ····················
G 9252 hCoV-19/Wales/PHWC-169BA9/2020|EPI_ISL_517599|2020 ···················· ···················· ····················· ···················· ···················· ····················
G 9253 hCoV-19/Suriname/SR-57/2020|EPI_ISL_518806|2020-07 ···················· ···················· ····················· ···················· ···················· ····················
G 9254 hCoV-19/Suriname/SR-58/2020|EPI_ISL_518807|2020-07 ···················· ···················· ····················· ···················· ···················· ····················
G 9255 hCoV-19/Suriname/SR-59/2020|EPI_ISL_518808|2020-07 ···················· ···················· ····················· ···················· ···················· ····················
G 9256 hCoV-19/Suriname/SR-60/2020|EPI_ISL_518809|2020-07 ···················· ···················· ····················· ···················· ···················· ····················
G 9257 hCoV-19/Suriname/SR-61/2020|EPI_ISL_518810|2020-07 ···················· ···················· ····················· ···················· ···················· ····················
G 9258 hCoV-19/Suriname/SR-63/2020|EPI_ISL_518812|2020-07 ···················· ···················· ····················· ···················· ···················· ····················
G 9259 hCoV-19/Suriname/SR-65/2020|EPI_ISL_518814|2020-07 ···············A···· ···················· ····················· ···················· ···················· ····················
G 9260 hCoV-19/Suriname/SR-67/2020|EPI_ISL_518816|2020-07 ···············A···· ···················· ····················· ···················· ···················· ····················
G 9261 hCoV-19/Suriname/SR-68/2020|EPI_ISL_518817|2020-07 ···············A···· ···················· ····················· ···················· ···················· ····················
G 9262 hCoV-19/Indonesia/TNG-LIPI001/2020|EPI_ISL_518819| ···················· ···················· ····················· ···················· ···················· ····················

```
G    9263 hCoV-19/Oman/RESP-20-16211/2020|EPI_ISL_518821|202     ····················  ·A··················  ····················  ····················  ····················  ····················
G    9264 hCoV-19/Oman/RESP-20-4395/2020|EPI_ISL_518822|2020     ····················  ····················  ····················  ····················  ····················  ····················
G    9265 hCoV-19/Oman/RESP-20-4941/2020|EPI_ISL_518823|2020     ····················  ····················  ····················  ····················  ····················  ····················
G    9266 hCoV-19/Oman/RESP-20-B-15095/2020|EPI_ISL_518824|2     ····················  ····················  ····················  ····················  ····················  ····················
G    9267 hCoV-19/Oman/RESP-20-B-15133/2020|EPI_ISL_518825|2     ····················  ····················  ····················  ····················  ····················  ····················
G    9268 hCoV-19/Oman/RESP-20-B-15173/2020|EPI_ISL_518826|2     ····················  ····················  ····················  ····················  ····················  ····················
G    9269 hCoV-19/Oman/RESP-20-B-15176/2020|EPI_ISL_518827|2     ····················  ····················  ····················  ····················  ····················  ····················
G    9270 hCoV-19/Oman/RESP-20-B-15181/2020|EPI_ISL_518828|2     ····················  ····················  ····················  ····················  ····················  ····················
G    9271 hCoV-19/Oman/RESP-20-B-15345/2020|EPI_ISL_518829|2     ····················  ····················  ····················  ····················  ····················  ····················
G    9272 hCoV-19/Oman/RESP-20-B-15411/2020|EPI_ISL_518831|2     ····················  ····················  ····················  ····················  ····················  ····················
G    9273 hCoV-19/Oman/RESP-20-B-15733/2020|EPI_ISL_518833|2     ····················  ····················  ····················  ····················  ····················  ····················
G    9274 hCoV-19/Oman/RESP-20-B-15753/2020|EPI_ISL_518834|2     ····················  ····················  ····················  ····················  ····················  ····················
G    9275 hCoV-19/Oman/RESP-20-B-15879/2020|EPI_ISL_518835|2     ····················  ····················  ····················  ····················  ····················  ····················
G    9276 hCoV-19/Oman/RESP-20-B-15905/2020|EPI_ISL_518836|2     ····················  ····················  ····················  ····················  ····················  ····················
G    9277 hCoV-19/Oman/RESP-20-B-16232/2020|EPI_ISL_518837|2     ····················  ····················  ····················  ····················  ····················  ····················
G    9278 hCoV-19/Oman/RESP-20-B-16236/2020|EPI_ISL_518838|2     ····················  ····················  ····················  ····················  ····················  ····················
G    9279 hCoV-19/Oman/RESP-20-B-16254/2020|EPI_ISL_518840|2     ····················  ····················  ····················  ····················  ····················  ····················
G    9280 hCoV-19/Oman/RESP-20-B-16294/2020|EPI_ISL_518841|2     ····················  ····················  ····················  ····················  ····················  ····················
G    9281 hCoV-19/USA/MN-MDH-1497/2020|EPI_ISL_518890|2020-0     ····················  ····················  ····················  ····················  ····················  ····················
G    9282 hCoV-19/USA/MN-MDH-1498/2020|EPI_ISL_518891|2020-0     ····················  ····················  ····················  ····················  ····················  ····················
G    9283 hCoV-19/USA/MN-MDH-1499/2020|EPI_ISL_518892|2020-0     ····················  ····················  ····················  ····················  ····················  ····················
G    9284 hCoV-19/USA/MN-MDH-1500/2020|EPI_ISL_518893|2020-0     ····················  ····················  ····················  ····················  ····················  ····················
G    9285 hCoV-19/USA/MN-MDH-1501/2020|EPI_ISL_518894|2020-0     ····················  ····················  ····················  ····················  ····················  ····················
G    9286 hCoV-19/USA/MN-MDH-1502/2020|EPI_ISL_518895|2020-0     ····················  ····················  ····················  ····················  ····················  ····················
G    9287 hCoV-19/USA/MN-MDH-1503/2020|EPI_ISL_518896|2020-0     ····················  ····················  ····················  ····················  ····················  ····················
G    9288 hCoV-19/USA/MN-MDH-1504/2020|EPI_ISL_518897|2020-0     ····················  ····················  ····················  ····················  ·····T··············  ····················
G    9289 hCoV-19/USA/MN-MDH-1505/2020|EPI_ISL_518898|2020-0     ····················  ····················  ····················  ····················  ····················  ····················
G    9290 hCoV-19/USA/MN-MDH-1506/2020|EPI_ISL_518899|2020-0     ····················  ····················  ····················  ····················  ····················  ····················
G    9291 hCoV-19/Australia/VIC2350/2020|EPI_ISL_522070|2020     ····················  ····················  ····················  ····················  ····················  ····················
G    9292 hCoV-19/Australia/VIC2351/2020|EPI_ISL_522071|2020     ····················  ····················  ····················  ····················  ····················  ····················
G    9293 hCoV-19/Australia/VIC2352/2020|EPI_ISL_522072|2020     ····················  ····················  ····················  ····················  NNN·················  ····················          1
G    9294 hCoV-19/Australia/VIC2353/2020|EPI_ISL_522073|2020     ····················  ····················  ····················  ····················  ····················  ····················
G    9296 hCoV-19/Australia/VIC2355/2020|EPI_ISL_522075|2020     ····················  ····················  ····················  ····················  ····················  ····················
G    9297 hCoV-19/Australia/VIC2357/2020|EPI_ISL_522077|2020     ····················  ····················  ····················  ····················  ····················  ····················
G    9298 hCoV-19/Australia/VIC2358/2020|EPI_ISL_522078|2020     ····················  ····················  ····················  ····················  ····················  ····················
G    9299 hCoV-19/Australia/VIC2359/2020|EPI_ISL_522079|2020     ····················  ····················  ····················  ····················  ····················  ····················
G    9300 hCoV-19/Australia/VIC2360/2020|EPI_ISL_522080|2020     ····················  ····················  ····················  ····················  ····················  ····················
G    9302 hCoV-19/Australia/VIC2362/2020|EPI_ISL_522082|2020     ····················  ····················  ····················  ····················  ····················  ····················
G    9303 hCoV-19/Australia/VIC2363/2020|EPI_ISL_522083|2020     ····················  ····················  ····················  ····················  ····················  ····················
G    9304 hCoV-19/Australia/VIC2364/2020|EPI_ISL_522084|2020     ····················  ····················  ····················  ····················  ····················  ····················
G    9305 hCoV-19/Australia/VIC2365/2020|EPI_ISL_522085|2020     ····················  ····················  ····················  ····················  ····················  ····················
G    9308 hCoV-19/Australia/VIC2368/2020|EPI_ISL_522088|2020     ····················  ····················  ····················  ····················  ····················  ····················
G    9309 hCoV-19/Australia/VIC2369/2020|EPI_ISL_522089|2020     ····················  ····················  ····················  ····················  ····················  ····················
G    9310 hCoV-19/Australia/VIC2371/2020|EPI_ISL_522091|2020     ····················  ····················  ····················  ····················  ····················  ····················
G    9311 hCoV-19/Australia/VIC2373/2020|EPI_ISL_522093|2020     ····················  ····················  ····················  ····················  ····················  ····················
G    9312 hCoV-19/Australia/VIC2374/2020|EPI_ISL_522094|2020     ····················  ····················  ····················  ····················  ····················  ····················
G    9313 hCoV-19/Australia/VIC2376/2020|EPI_ISL_522096|2020     ····················  ····················  ····················  ····················  ····················  ····················
G    9314 hCoV-19/Australia/VIC2377/2020|EPI_ISL_522097|2020     ····················  ····················  ····················  ····················  ····················  ····················
G    9315 hCoV-19/Australia/VIC2378/2020|EPI_ISL_522098|2020     ····················  ····················  ····················  ····················  ····················  ····················
G    9317 hCoV-19/Switzerland/BL-UHB-42197274/2020|EPI_ISL_5     ····················  ····················  ····················  ····················  ····················  ····················
G    9318 hCoV-19/Switzerland/BS-UHB-42197347/2020|EPI_ISL_5     ····················  ····················  ····················  ····················  ····················  ····················
G    9319 hCoV-19/Switzerland/BS-UHB-42197488/2020|EPI_ISL_5     ····················  ····················  ····················  ····················  ····················  ····················
G    9320 hCoV-19/Switzerland/BL-UHB-42197574/2020|EPI_ISL_5     ····················  ····················  ····················  ····················  ····················  ····················
G    9321 hCoV-19/Switzerland/BS-UHB-42197601/2020|EPI_ISL_5     ····················  ····················  ····················  ····················  ····················  ····················
G    9322 hCoV-19/Switzerland/BS-UHB-42197732/2020|EPI_ISL_5     ····················  ····················  ····················  ····················  ····················  ····················
G    9324 hCoV-19/Switzerland/BS-UHB-42198593/2020|EPI_ISL_5     ····················  ····················  ····················  ····················  ····················  ····················
G    9325 hCoV-19/Switzerland/BS-UHB-42198617/2020|EPI_ISL_5     ····················  ····················  ····················  ····················  ····················  ····················
G    9326 hCoV-19/Switzerland/BL-UHB-42198619/2020|EPI_ISL_5     ····················  ···················A  ····················  ····················  ····················  ····················       1
G    9327 hCoV-19/Switzerland/BL-UHB-42198620/2020|EPI_ISL_5     ····················  ···················A  ····················  ····················  ····················  ····················       1
G    9328 hCoV-19/Switzerland/BS-UHB-42198664/2020|EPI_ISL_5     ····················  ····················  ····················  ····················  ····················  ····················
G    9329 hCoV-19/Switzerland/BS-UHB-42198756/2020|EPI_ISL_5     ····················  ····················  ····················  ····················  ····················  ····················
G    9330 hCoV-19/Switzerland/BL-UHB-42198812/2020|EPI_ISL_5     ····················  ····················  ····················  ····················  ····················  ····················
G    9331 hCoV-19/Switzerland/BS-UHB-42198823/2020|EPI_ISL_5     ····················  ····················  ····················  ····················  ····················  ····················
G    9332 hCoV-19/Switzerland/BS-UHB-42198935/2020|EPI_ISL_5     ····················  ····················  ····················  ····················  ····················  ····················
G    9333 hCoV-19/Switzerland/BS-UHB-42199036/2020|EPI_ISL_5     ····················  ····················  ····················  ····················  ····················  ····················
G    9334 hCoV-19/Switzerland/BS-UHB-42199076/2020|EPI_ISL_5     ····················  ····················  ····················  ····················  ····················  ····················
G    9335 hCoV-19/Switzerland/BS-UHB-42199107/2020|EPI_ISL_5     ····················  ····················  ····················  ····················  ····················  ····················
G    9336 hCoV-19/Switzerland/BS-UHB-42199265/2020|EPI_ISL_5     ····················  ····················  ····················  ····················  ····················  ····················
G    9337 hCoV-19/Switzerland/BS-UHB-42199640/2020|EPI_ISL_5     ····················  ····················  ····················  ····················  ····················  ····················
G    9338 hCoV-19/Switzerland/BS-UHB-42199675/2020|EPI_ISL_5     ····················  ····················  ····················  ····················  ····················  ····················
G    9339 hCoV-19/Switzerland/BS-UHB-42199704/2020|EPI_ISL_5     ····················  ····················  ····················  ····················  ····················  ····················
G    9340 hCoV-19/Switzerland/BS-UHB-42199724/2020|EPI_ISL_5     ····················  ····················  ····················  ····················  ····················  ····················
G    9341 hCoV-19/Switzerland/BL-UHB-42199737/2020|EPI_ISL_5     ····················  ····················  ····················  ····················  ····················  ····················
G    9342 hCoV-19/Switzerland/BS-UHB-42199741/2020|EPI_ISL_5     ····················  ····················  ····················  ····················  ····················  ····················
G    9343 hCoV-19/Switzerland/BS-UHB-42199777/2020|EPI_ISL_5     ····················  ····················  ····················  ····················  ····················  ····················
G    9344 hCoV-19/Switzerland/BS-UHB-42199797/2020|EPI_ISL_5     ····················  ····················  ····················  ····················  ····················  ····················
G    9345 hCoV-19/Switzerland/BS-UHB-42199808/2020|EPI_ISL_5     ····················  ····················  ····················  ····················  ····················  ····················
G    9346 hCoV-19/Switzerland/BS-UHB-42199841/2020|EPI_ISL_5     ····················  ····················  ····················  ····················  ····················  ····················
G    9347 hCoV-19/Switzerland/BS-UHB-42199864/2020|EPI_ISL_5     ····················  ····················  ····················  ····················  ····················  ····················
G    9348 hCoV-19/Switzerland/BS-UHB-42199899/2020|EPI_ISL_5     ····················  ····················  ····················  ····················  ····················  ····················
```

| | | | | | | | | |
|---|---|---|---|---|---|---|---|---|
| G | 9349 hCoV-19/Switzerland/BS-UHB-42199905/2020\|EPI_ISL_5 | .................... | .................... | .................... | .................... | .................... | .................... | |
| G | 9350 hCoV-19/Switzerland/BS-UHB-42199917/2020\|EPI_ISL_5 | .................... | .................... | .................... | .................... | .................... | .................... | |
| G | 9351 hCoV-19/Switzerland/BS-UHB-42199930/2020\|EPI_ISL_5 | .................... | .................... | .................... | .................... | .................... | .................... | |
| G | 9352 hCoV-19/Switzerland/BS-UHB-42199957/2020\|EPI_ISL_5 | .................... | .................... | .................... | .................... | .................... | .................... | |
| G | 9353 hCoV-19/Switzerland/BS-UHB-42200029/2020\|EPI_ISL_5 | .................... | .A.................. | .................... | .................... | .................... | .................... | |
| G | 9354 hCoV-19/Switzerland/BS-UHB-42200048/2020\|EPI_ISL_5 | .................... | .................... | .................... | .................... | .................... | .................... | |
| G | 9355 hCoV-19/Switzerland/BL-UHB-42200078/2020\|EPI_ISL_5 | .................... | .................... | .................... | .................... | .................... | .................... | |
| G | 9356 hCoV-19/Switzerland/BL-UHB-42200091/2020\|EPI_ISL_5 | .................... | .................... | .................... | .................... | .................... | .................... | |
| G | 9357 hCoV-19/Switzerland/BS-UHB-42200118/2020\|EPI_ISL_5 | .................... | .................... | .................... | .................... | .................... | .................... | |
| G | 9358 hCoV-19/Switzerland/BS-UHB-42200131/2020\|EPI_ISL_5 | .................... | .................... | .................... | .................... | .................... | .................... | |
| G | 9359 hCoV-19/Switzerland/BS-UHB-42200134/2020\|EPI_ISL_5 | .................... | .A.................. | .................... | .................... | .................... | .................... | |
| G | 9360 hCoV-19/Switzerland/BS-UHB-42200181/2020\|EPI_ISL_5 | .................... | .................... | .................... | .................... | .................... | .................... | |
| G | 9361 hCoV-19/Switzerland/BS-UHB-42200253/2020\|EPI_ISL_5 | .................... | .................... | .................... | .................... | .................... | .................... | |
| G | 9362 hCoV-19/Switzerland/BS-UHB-42200960/2020\|EPI_ISL_5 | .................... | .................... | .................... | .................... | .................... | .................... | |
| G | 9363 hCoV-19/Switzerland/BL-UHB-42200964/2020\|EPI_ISL_5 | .................... | .................... | .................... | .................... | .................... | .................... | |
| G | 9364 hCoV-19/Switzerland/BS-UHB-42200965/2020\|EPI_ISL_5 | .................... | .................... | .................... | .................... | .................... | .................... | |
| G | 9365 hCoV-19/Switzerland/BS-UHB-42201016/2020\|EPI_ISL_5 | .................... | .................... | .................... | .................... | .................... | .................... | |
| G | 9366 hCoV-19/Switzerland/BS-UHB-42201233/2020\|EPI_ISL_5 | .................... | .................... | .................... | .................... | .................... | .................... | |
| G | 9367 hCoV-19/Switzerland/BS-UHB-42201257/2020\|EPI_ISL_5 | .................... | .................... | .................... | .................... | .................... | .................... | |
| G | 9368 hCoV-19/Switzerland/BS-UHB-42201258/2020\|EPI_ISL_5 | .................... | .................... | .................... | .................... | .................... | .................... | |
| G | 9369 hCoV-19/Switzerland/BS-UHB-42201259/2020\|EPI_ISL_5 | .................... | .................... | .................... | .................... | .................... | .................... | |
| G | 9370 hCoV-19/Switzerland/BS-UHB-42201261/2020\|EPI_ISL_5 | .................... | ...................A | .................... | .................... | .................... | .................... | 1 |
| G | 9371 hCoV-19/Switzerland/BS-UHB-42201263/2020\|EPI_ISL_5 | .................... | ...................A | .................... | .................... | .................... | .................... | 1 |
| G | 9372 hCoV-19/England/ALDP-9805C4/2020\|EPI_ISL_529540\|20 | .................... | .................... | .................... | .................... | .................... | .................... | |
| G | 9373 hCoV-19/England/ALDP-980B59/2020\|EPI_ISL_529541\|20 | .................... | .................... | .................... | .................... | .................... | .................... | |
| G | 9374 hCoV-19/England/ALDP-980B68/2020\|EPI_ISL_529542\|20 | .................... | .A.................. | .................... | .................... | .................... | ..A................. | |
| G | 9375 hCoV-19/England/ALDP-980B77/2020\|EPI_ISL_529543\|20 | .................... | .................... | .................... | .................... | .................... | .................... | |
| G | 9376 hCoV-19/England/ALDP-980F02/2020\|EPI_ISL_529544\|20 | .................... | .A.................. | .................... | .................... | .................... | ..A................. | |
| G | 9377 hCoV-19/England/ALDP-9813C9/2020\|EPI_ISL_529545\|20 | .................... | .................... | .................... | .................... | .................... | .................... | |
| G | 9378 hCoV-19/England/ALDP-981666/2020\|EPI_ISL_529546\|20 | .................... | .................... | .................... | .................... | .................... | .................... | |
| G | 9379 hCoV-19/England/ALDP-981CA0/2020\|EPI_ISL_529547\|20 | .................... | .................... | .................... | .................... | .................... | .................... | |
| G | 9380 hCoV-19/England/ALDP-981E7D/2020\|EPI_ISL_529548\|20 | .................... | .A.................. | .................... | .................... | .................... | ..A................. | |
| G | 9381 hCoV-19/England/ALDP-981FA7/2020\|EPI_ISL_529549\|20 | .................... | .................... | .................... | .................... | .................... | .................... | |
| G | 9382 hCoV-19/England/ALDP-982674/2020\|EPI_ISL_529550\|20 | .................... | .A.................. | .................... | .................... | .................... | ..A................. | |
| G | 9383 hCoV-19/England/ALDP-9827EA/2020\|EPI_ISL_529551\|20 | .................... | .................... | .................... | .................... | .................... | .................... | |
| G | 9384 hCoV-19/England/ALDP-982AB4/2020\|EPI_ISL_529552\|20 | .................... | .................... | .................... | .................... | .................... | .................... | |
| G | 9385 hCoV-19/England/ALDP-983215/2020\|EPI_ISL_529553\|20 | .................... | .................... | .................... | .................... | .................... | .................... | |
| G | 9386 hCoV-19/England/ALDP-9832F7/2020\|EPI_ISL_529554\|20 | .................... | .................... | .................... | .................... | .................... | .................... | |
| G | 9387 hCoV-19/England/ALDP-983303/2020\|EPI_ISL_529555\|20 | .................... | .................... | .................... | .................... | .................... | .................... | |
| G | 9388 hCoV-19/England/ALDP-983576/2020\|EPI_ISL_529556\|20 | .................... | .................... | .................... | .................... | .................... | .................... | |
| G | 9389 hCoV-19/England/ALDP-983822/2020\|EPI_ISL_529557\|20 | .................... | .................... | .................... | .................... | .................... | .................... | |
| G | 9390 hCoV-19/England/ALDP-98393E/2020\|EPI_ISL_529558\|20 | .................... | .................... | .................... | .................... | .................... | .................... | |
| G | 9391 hCoV-19/England/ALDP-98394D/2020\|EPI_ISL_529559\|20 | .................... | .................... | .................... | .................... | .................... | .................... | |
| G | 9392 hCoV-19/England/ALDP-9842D8/2020\|EPI_ISL_529560\|20 | .................... | .................... | .................... | .................... | .................... | .................... | |
| G | 9393 hCoV-19/England/ALDP-9846AF/2020\|EPI_ISL_529561\|20 | .................... | .................... | .................... | .................... | .................... | .................... | |
| G | 9394 hCoV-19/England/ALDP-9846BE/2020\|EPI_ISL_529562\|20 | .................... | .................... | .................... | .................... | .................... | .................... | |
| G | 9395 hCoV-19/England/ALDP-9846CD/2020\|EPI_ISL_529563\|20 | .................... | .................... | .................... | .................... | .................... | .................... | |
| G | 9396 hCoV-19/England/ALDP-9846DC/2020\|EPI_ISL_529564\|20 | .................... | .................... | .................... | .................... | .................... | .................... | |
| G | 9397 hCoV-19/England/ALDP-9846EB/2020\|EPI_ISL_529565\|20 | .................... | .................... | .................... | .................... | .................... | .................... | |
| G | 9398 hCoV-19/England/ALDP-9846FA/2020\|EPI_ISL_529566\|20 | .................... | .................... | .................... | .................... | .................... | .................... | |
| G | 9399 hCoV-19/England/ALDP-984C43/2020\|EPI_ISL_529567\|20 | .................... | .................... | .................... | .................... | .................... | .................... | |
| G | 9400 hCoV-19/England/ALDP-984D04/2020\|EPI_ISL_529568\|20 | .................... | .................... | .................... | .................... | .................... | .................... | |
| G | 9401 hCoV-19/England/ALDP-984FA4/2020\|EPI_ISL_529569\|20 | .................... | .................... | .................... | .................... | .................... | .................... | |
| G | 9402 hCoV-19/England/ALDP-9859E1/2020\|EPI_ISL_529570\|20 | .................... | .................... | .................... | .................... | .................... | .................... | |
| G | 9403 hCoV-19/England/ALDP-985A57/2020\|EPI_ISL_529571\|20 | .................... | .................... | .................... | .................... | .................... | .................... | |
| G | 9404 hCoV-19/England/ALDP-985BDC/2020\|EPI_ISL_529572\|20 | .................... | .................... | .................... | .................... | .................... | .................... | |
| G | 9405 hCoV-19/England/ALDP-98634C/2020\|EPI_ISL_529573\|20 | .................... | .A.................. | .................... | .................... | .................... | ..A................. | |
| G | 9406 hCoV-19/England/ALDP-9863D3/2020\|EPI_ISL_529574\|20 | .................... | .................... | .................... | .................... | .................... | .................... | |
| G | 9407 hCoV-19/England/ALDP-9863F1/2020\|EPI_ISL_529575\|20 | .................... | .................... | .................... | .................... | .................... | .................... | |
| G | 9408 hCoV-19/England/ALDP-986731/2020\|EPI_ISL_529576\|20 | .................... | .A.................. | .................... | .................... | .................... | ..A................. | |
| G | 9409 hCoV-19/England/ALDP-98733C/2020\|EPI_ISL_529577\|20 | .................... | .A.................. | .................... | .................... | .................... | ..A................. | |
| G | 9410 hCoV-19/England/ALDP-987651/2020\|EPI_ISL_529578\|20 | .................... | .................... | .................... | .................... | .................... | .................... | |
| G | 9411 hCoV-19/England/ALDP-987703/2020\|EPI_ISL_529579\|20 | .................... | .................... | .................... | .................... | .................... | .................... | |
| G | 9412 hCoV-19/England/ALDP-987712/2020\|EPI_ISL_529580\|20 | .................... | .................... | .................... | .................... | .................... | .................... | |
| G | 9413 hCoV-19/England/ALDP-9877A9/2020\|EPI_ISL_529581\|20 | .................... | .................... | .................... | .................... | .................... | .................... | |
| G | 9414 hCoV-19/England/ALDP-987E2C/2020\|EPI_ISL_529582\|20 | .................... | .................... | .................... | .................... | .................... | .................... | |
| G | 9415 hCoV-19/England/ALDP-987E3B/2020\|EPI_ISL_529583\|20 | .................... | .................... | .................... | .................... | .................... | .................... | |
| G | 9416 hCoV-19/England/ALDP-987E86/2020\|EPI_ISL_529584\|20 | .................... | .................... | .................... | .................... | .................... | .................... | |
| G | 9417 hCoV-19/England/ALDP-987EB3/2020\|EPI_ISL_529585\|20 | .................... | .................... | .................... | .................... | .................... | .................... | |
| G | 9418 hCoV-19/England/ALDP-987ED1/2020\|EPI_ISL_529586\|20 | .................... | .................... | .................... | .................... | .................... | .................... | |
| G | 9419 hCoV-19/England/ALDP-988483/2020\|EPI_ISL_529587\|20 | .................... | .................... | .................... | .................... | .................... | .................... | |
| G | 9420 hCoV-19/England/ALDP-9884B0/2020\|EPI_ISL_529588\|20 | .................... | .A.................. | .................... | .................... | .................... | ..A................. | |
| G | 9421 hCoV-19/England/ALDP-988C4F/2020\|EPI_ISL_529589\|20 | .................... | .................... | .................... | .................... | .................... | .................... | |
| G | 9422 hCoV-19/England/BIRM-60CFB/2020\|EPI_ISL_529590\|202 | .................... | .................... | .................... | .................... | .................... | .................... | |
| G | 9423 hCoV-19/England/BIRM-60DCB/2020\|EPI_ISL_529591\|202 | .................... | .................... | .................... | .................... | .................... | .................... | |
| G | 9424 hCoV-19/England/BIRM-60DE9/2020\|EPI_ISL_529592\|202 | .................... | .................... | .................... | .................... | .................... | .................... | |
| G | 9425 hCoV-19/England/BIRM-60E04/2020\|EPI_ISL_529593\|202 | .................... | .................... | .................... | .................... | .................... | .................... | |
| G | 9426 hCoV-19/England/BIRM-60E13/2020\|EPI_ISL_529594\|202 | .................... | .................... | .................... | .................... | .................... | .................... | |
| G | 9427 hCoV-19/England/BIRM-60E7D/2020\|EPI_ISL_529595\|202 | .................... | .................... | .................... | .................... | .................... | .................... | |
| G | 9428 hCoV-19/England/BIRM-6189C/2020\|EPI_ISL_529596\|202 | .................... | .................... | .................... | .................... | .................... | .................... | |

| | | | | | | | | | |
|---|---|---|---|---|---|---|---|---|---|
| G | 9429 | hCoV-19/England/BIRM-61A87/2020\|EPI_ISL_529597\|202 | ···················· | ···················· | ···················· | ···················· | ···················· | ···················· | |
| G | 9430 | hCoV-19/England/BIRM-61B39/2020\|EPI_ISL_529598\|202 | ···················· | ···················· | ···················· | ···················· | ···················· | ···················· | |
| G | 9431 | hCoV-19/England/BIRM-61B75/2020\|EPI_ISL_529599\|202 | ···················· | ···················· | ···················· | ···················· | ···················· | ···················· | |
| H | 1 | hCoV-19/England/QEUH-9606A9/2020\|EPI_ISL_532101\|20 | ···················· | ···················· | ···················· | ···················· | ···················· | ···················· | |
| H | 2 | hCoV-19/Scotland/QEUH-9611D4/2020\|EPI_ISL_532102\|2 | ···················· | ···················· | ···················· | ···················· | ···················· | ···················· | |
| H | 3 | hCoV-19/England/QEUH-960E1A/2020\|EPI_ISL_532103\|20 | ···················· | ·A·················· | ···················· | ···················· | ···················· | ···················· | |
| H | 4 | hCoV-19/England/QEUH-960867/2020\|EPI_ISL_532104\|20 | ···················· | ···················· | ···················· | ···················· | ···················· | ···················· | |
| H | 5 | hCoV-19/Scotland/QEUH-960463/2020\|EPI_ISL_532105\|2 | ···················· | ···················· | ···················· | ···················· | ···················· | ···················· | |
| H | 6 | hCoV-19/England/QEUH-961022/2020\|EPI_ISL_532106\|20 | ···················· | ···················· | ···················· | ···················· | ···················· | ···················· | |
| H | 7 | hCoV-19/England/QEUH-960EED/2020\|EPI_ISL_532107\|20 | ···················· | ···················· | ···················· | ···················· | ···················· | ···················· | |
| H | 8 | hCoV-19/England/QEUH-960C5C/2020\|EPI_ISL_532109\|20 | ···················· | ·A·················· | ···················· | ···················· | ···················· | ··A················· | |
| H | 9 | hCoV-19/England/QEUH-9612E0/2020\|EPI_ISL_532110\|20 | ···················· | ···················· | ···················· | ···················· | ···················· | ···················· | |
| H | 10 | hCoV-19/England/QEUH-960366/2020\|EPI_ISL_532111\|20 | ···················· | ···················· | ···················· | ···················· | ···················· | ···················· | |
| H | 11 | hCoV-19/England/QEUH-960B31/2020\|EPI_ISL_532112\|20 | ···················· | ···················· | ···················· | ···················· | ···················· | ···················· | |
| H | 12 | hCoV-19/England/QEUH-961338/2020\|EPI_ISL_532113\|20 | ···················· | ···················· | ···················· | ···················· | ···················· | ···················· | |
| H | 13 | hCoV-19/England/QEUH-960375/2020\|EPI_ISL_532114\|20 | ···················· | ···················· | ···················· | ···················· | ···················· | ···················· | |
| H | 14 | hCoV-19/England/QEUH-9607A6/2020\|EPI_ISL_532115\|20 | ···················· | ···················· | ···················· | ···················· | ···················· | ···················· | |
| H | 15 | hCoV-19/England/QEUH-96090A/2020\|EPI_ISL_532116\|20 | ···················· | ···················· | ···················· | ···················· | ···················· | ···················· | |
| H | 16 | hCoV-19/England/QEUH-96083A/2020\|EPI_ISL_532117\|20 | ···················· | ···················· | ···················· | ···················· | ···················· | ···················· | |
| H | 17 | hCoV-19/Scotland/QEUH-9611F2/2020\|EPI_ISL_532118\|2 | ···················· | ·A·················· | ···················· | ···················· | ···················· | ···················· | |
| H | 18 | hCoV-19/England/QEUH-960427/2020\|EPI_ISL_532119\|20 | ···················· | ···················· | ···················· | ···················· | ···················· | ···················· | |
| H | 19 | hCoV-19/England/QEUH-960E38/2020\|EPI_ISL_532120\|20 | ···················· | ···················· | ···················· | ···················· | ···················· | ···················· | |
| H | 20 | hCoV-19/England/QEUH-960630/2020\|EPI_ISL_532121\|20 | ···················· | ···················· | ···················· | ···················· | ···················· | ···················· | |
| H | 21 | hCoV-19/England/QEUH-960AAD/2020\|EPI_ISL_532122\|20 | ···················· | ···················· | ···················· | ···················· | ···················· | ···················· | |
| H | 22 | hCoV-19/Scotland/QEUH-96073D/2020\|EPI_ISL_532123\|2 | ···················· | ···················· | ···················· | ···················· | ···················· | ···················· | |
| H | 23 | hCoV-19/England/QEUH-960490/2020\|EPI_ISL_532124\|20 | ···················· | ···················· | ···················· | ···················· | ···················· | ···················· | |
| H | 24 | hCoV-19/Scotland/QEUH-96116B/2020\|EPI_ISL_532125\|2 | ···················· | ···················· | ···················· | ···················· | ···················· | ···················· | |
| H | 25 | hCoV-19/England/QEUH-960928/2020\|EPI_ISL_532126\|20 | ···················· | ···················· | ···················· | ···················· | ···················· | ···················· | |
| H | 26 | hCoV-19/England/QEUH-960E29/2020\|EPI_ISL_532127\|20 | ···················· | ·A·················· | ···················· | ···················· | ···················· | ···················· | |
| H | 27 | hCoV-19/Northern Ireland/QEUH-960DB3/2020\|EPI_ISL_ | ···················· | ···················· | ···················· | ···················· | ···················· | ···················· | |
| H | 28 | hCoV-19/Scotland/QEUH-96112F/2020\|EPI_ISL_532129\|2 | ···················· | ···················· | ···················· | ···················· | ···················· | ···················· | |
| H | 29 | hCoV-19/Scotland/QEUH-9604AF/2020\|EPI_ISL_532130\|2 | ···················· | ···················· | ···················· | ···················· | ···················· | ···················· | |
| H | 30 | hCoV-19/Scotland/QEUH-96117A/2020\|EPI_ISL_532131\|2 | ···················· | ···················· | ···················· | ···················· | ···················· | ···················· | |
| H | 31 | hCoV-19/England/QEUH-96066D/2020\|EPI_ISL_532132\|20 | ···················· | ·A······CA·········· | ···················· | ···················· | ···················· | ···················· | 1 |
| H | 32 | hCoV-19/England/QEUH-960ABC/2020\|EPI_ISL_532133\|20 | ···················· | ···················· | ···················· | ···················· | ···················· | ···················· | |
| H | 33 | hCoV-19/England/QEUH-9608A3/2020\|EPI_ISL_532134\|20 | ···················· | ···················· | ···················· | ···················· | ···················· | ···················· | |
| H | 34 | hCoV-19/England/QEUH-960F17/2020\|EPI_ISL_532135\|20 | ···················· | ·A·················· | ···················· | ···················· | ···················· | ···················· | |
| H | 35 | hCoV-19/England/QEUH-960CD4/2020\|EPI_ISL_532136\|20 | ···················· | ·A·················· | ···················· | ···················· | ···················· | ···················· | |
| H | 36 | hCoV-19/England/QEUH-9605D9/2020\|EPI_ISL_532137\|20 | ···················· | ···················· | ···················· | ···················· | ······T············· | ···················· | |
| H | 37 | hCoV-19/Scotland/QEUH-96120E/2020\|EPI_ISL_532138\|2 | ···················· | ···················· | ···················· | ···················· | ···················· | ···················· | |
| H | 38 | hCoV-19/England/QEUH-53C446/2020\|EPI_ISL_532140\|20 | ···················· | ···················· | ···················· | ···················· | ···················· | ···················· | |
| H | 39 | hCoV-19/England/QEUH-53F0B8/2020\|EPI_ISL_532141\|20 | ···················· | ···················· | ···················· | ···················· | ···················· | ···················· | |
| H | 40 | hCoV-19/England/QEUH-554C9B/2020\|EPI_ISL_532143\|20 | ···················· | ···················· | ···················· | ···················· | ···················· | ···················· | |
| H | 41 | hCoV-19/England/QEUH-55630D/2020\|EPI_ISL_532144\|20 | ···················· | ···················· | ···················· | ···················· | ···················· | ···················· | |
| H | 42 | hCoV-19/England/QEUH-539D6C/2020\|EPI_ISL_532146\|20 | ···················· | ···················· | ···················· | ···················· | ···················· | ···················· | |
| H | 43 | hCoV-19/England/QEUH-53BD4C/2020\|EPI_ISL_532147\|20 | ···················· | ···················· | ···················· | ···················· | ···················· | ···················· | |
| H | 44 | hCoV-19/England/QEUH-557041/2020\|EPI_ISL_532148\|20 | ···················· | ···················· | ···················· | ···················· | ···················· | ···················· | |
| H | 45 | hCoV-19/England/QEUH-52DBE9/2020\|EPI_ISL_532150\|20 | ···················· | ···················· | ···················· | ···················· | ···················· | ···················· | |
| H | 46 | hCoV-19/England/QEUH-5318CD/2020\|EPI_ISL_532151\|20 | ···················· | ···················· | ···················· | ···················· | ···················· | ···················· | |
| H | 47 | hCoV-19/England/QEUH-538EE2/2020\|EPI_ISL_532152\|20 | ···················· | ···················· | ···················· | ···················· | ···················· | ···················· | |
| H | 48 | hCoV-19/England/QEUH-557DC2/2020\|EPI_ISL_532156\|20 | ···················· | ···················· | ···················· | ···················· | ···················· | ···················· | |
| H | 49 | hCoV-19/England/QEUH-55336A/2020\|EPI_ISL_532157\|20 | ···················· | ···················· | ···················· | ···················· | ···················· | ···················· | |
| H | 50 | hCoV-19/England/QEUH-53D17B/2020\|EPI_ISL_532158\|20 | ···················· | ···················· | ···················· | ···················· | ···················· | ···················· | |
| H | 51 | hCoV-19/England/QEUH-53983E/2020\|EPI_ISL_532159\|20 | ···················· | ···················· | ···················· | ···················· | ···················· | ···················· | |
| H | 52 | hCoV-19/England/QEUH-53DABC/2020\|EPI_ISL_532160\|20 | ···················· | ···················· | ···················· | ···················· | ···················· | ···················· | |
| H | 53 | hCoV-19/England/QEUH-550DC9/2020\|EPI_ISL_532162\|20 | ···················· | ···················· | ···················· | ···················· | ···················· | ···················· | |
| H | 54 | hCoV-19/England/QEUH-554CB9/2020\|EPI_ISL_532164\|20 | ···················· | ···················· | ···················· | ···················· | ···················· | ···················· | |
| H | 55 | hCoV-19/England/QEUH-541EB9/2020\|EPI_ISL_532165\|20 | ···················· | ···················· | ···················· | ···················· | ···················· | ···················· | |
| H | 56 | hCoV-19/England/QEUH-5313BD/2020\|EPI_ISL_532166\|20 | ···················· | ···················· | ···················· | ···················· | ···················· | ···················· | |
| H | 57 | hCoV-19/England/QEUH-54E46F/2020\|EPI_ISL_532170\|20 | ···················· | ···················· | ···················· | ···················· | ···················· | ···················· | |
| H | 58 | hCoV-19/England/QEUH-53204C/2020\|EPI_ISL_532171\|20 | ···················· | ···················· | ···················· | ···················· | ···················· | ···················· | |
| H | 59 | hCoV-19/England/QEUH-55674D/2020\|EPI_ISL_532172\|20 | ···················· | ···················· | ···················· | ···················· | ···················· | ···················· | |
| H | 60 | hCoV-19/England/QEUH-53CB32/2020\|EPI_ISL_532173\|20 | ···················· | ···················· | ···················· | ···················· | ···················· | ···················· | |
| H | 61 | hCoV-19/England/QEUH-543DF6/2020\|EPI_ISL_532176\|20 | ···················· | ···················· | ···················· | ···················· | ···················· | ···················· | |
| H | 62 | hCoV-19/Scotland/QEUH-53D3A2/2020\|EPI_ISL_532178\|2 | ···················· | ···················· | ···················· | ···················· | ···················· | ···················· | |
| H | 63 | hCoV-19/England/QEUH-53A554/2020\|EPI_ISL_532180\|20 | ···················· | ···················· | ···················· | ···················· | ···················· | ···················· | |
| H | 64 | hCoV-19/England/QEUH-555BF7/2020\|EPI_ISL_532181\|20 | ···················· | ·A·················· | ···················· | ···················· | ···················· | ···················· | |
| H | 65 | hCoV-19/England/QEUH-555492/2020\|EPI_ISL_532183\|20 | ···················· | ···················· | ···················· | ···················· | ···················· | ···················· | |
| H | 66 | hCoV-19/England/QEUH-5528A8/2020\|EPI_ISL_532184\|20 | ···················· | ···················· | ···················· | ···················· | ···················· | ···················· | |
| H | 67 | hCoV-19/England/QEUH-52DACE/2020\|EPI_ISL_532185\|20 | ···················· | ·A·················· | ···················· | ···················· | ···················· | ···················· | |
| H | 68 | hCoV-19/England/QEUH-554D98/2020\|EPI_ISL_532188\|20 | ···················· | ···················· | ···················· | ···················· | ···················· | ···················· | |
| H | 69 | hCoV-19/England/QEUH-55484C/2020\|EPI_ISL_532190\|20 | ···················· | ···················· | ···················· | ···················· | ···················· | ···················· | |
| H | 70 | hCoV-19/England/QEUH-53E80C/2020\|EPI_ISL_532191\|20 | ···················· | ···················· | ···················· | ···················· | ···················· | ···················· | |
| H | 71 | hCoV-19/Scotland/QEUH-54542C/2020\|EPI_ISL_532192\|2 | ···················· | ···················· | ···················· | ···················· | ···················· | ···················· | |
| H | 72 | hCoV-19/England/QEUH-53C0AC/2020\|EPI_ISL_532193\|20 | ···················· | ···················· | ···················· | ···················· | ···················· | ···················· | |
| H | 73 | hCoV-19/Scotland/QEUH-53102F/2020\|EPI_ISL_532195\|2 | ···················· | ···················· | ···················· | ···················· | ···················· | ···················· | |
| H | 74 | hCoV-19/Scotland/QEUH-54EC3A/2020\|EPI_ISL_532196\|2 | ···················· | ···················· | ···················· | ···················· | ···················· | ···················· | |
| H | 75 | hCoV-19/England/QEUH-53E4AE/2020\|EPI_ISL_532198\|20 | ···················· | ···················· | ···················· | ···················· | ···················· | ···················· | |
| H | 76 | hCoV-19/England/QEUH-556F45/2020\|EPI_ISL_532199\|20 | ···················· | ···················· | ···················· | ···················· | ···················· | ···················· | |
| H | 77 | hCoV-19/England/QEUH-54A445/2020\|EPI_ISL_533400\|20 | ···················· | ···················· | ···················· | ···················· | ···················· | ···················· | |

| | | | | | | | | | |
|---|---|---|---|---|---|---|---|---|---|
| H | 78 | hCoV-19/England/QEUH-54724E/2020\|EPI_ISL_533401\|20 | ···················· | ···················· | ···················· | ···················· | ···················· | ···················· | |
| H | 79 | hCoV-19/England/QEUH-54D749/2020\|EPI_ISL_533402\|20 | ···················· | ···················· | ···················· | ···················· | ···················· | ···················· | |
| H | 80 | hCoV-19/England/QEUH-5494A0/2020\|EPI_ISL_533404\|20 | ···················· | ···················· | ···················· | ···················· | ···················· | ···················· | |
| H | 81 | hCoV-19/England/QEUH-5474EE/2020\|EPI_ISL_533405\|20 | ···················· | ···················· | ···················· | ···················· | ···················· | ···················· | |
| H | 82 | hCoV-19/Scotland/QEUH-549E48/2020\|EPI_ISL_533406\|2 | ···················· | ···················· | ···················· | ···················· | ···················· | ···················· | |
| H | 83 | hCoV-19/England/QEUH-5473D2/2020\|EPI_ISL_533407\|20 | ···················· | ···················· | ···················· | ···················· | ···················· | ···················· | |
| H | 84 | hCoV-19/England/QEUH-54AE47/2020\|EPI_ISL_533408\|20 | ···················· | ···················· | ···················· | ···················· | ···················· | ···················· | |
| H | 85 | hCoV-19/England/QEUH-547378/2020\|EPI_ISL_533409\|20 | ···················· | ···················· | ···················· | ···················· | ···················· | ···················· | |
| H | 86 | hCoV-19/England/QEUH-54B356/2020\|EPI_ISL_533410\|20 | ···················· | ···················A | ···················· | ···················· | ···················· | ···················· | 1 |
| H | 87 | hCoV-19/England/QEUH-54D55E/2020\|EPI_ISL_533412\|20 | ···················· | ···················· | ···················· | ···················· | ···················· | ···················· | |
| H | 88 | hCoV-19/Wales/QEUH-54C9CC/2020\|EPI_ISL_533413\|2020 | ···················· | ···················· | ···················· | ···················· | ···················· | ···················· | |
| H | 89 | hCoV-19/England/QEUH-548052/2020\|EPI_ISL_533414\|20 | ···················· | ···················· | ···················· | ···················· | ···················· | ···················· | |
| H | 90 | hCoV-19/England/QEUH-54BA06/2020\|EPI_ISL_533415\|20 | ···················· | ···················· | ···················· | ···················· | ···················· | ···················· | |
| H | 91 | hCoV-19/England/QEUH-548043/2020\|EPI_ISL_533416\|20 | ···················· | ···················· | ···················· | ···················· | ···················· | ···················· | |
| H | 92 | hCoV-19/England/QEUH-548CD6/2020\|EPI_ISL_533417\|20 | ···················· | ···················· | ···················· | ···················· | ···················· | ···················· | |
| H | 93 | hCoV-19/Wales/QEUH-54CB20/2020\|EPI_ISL_533418\|2020 | ···················· | ···················· | ···················· | ···················· | ···················· | ···················· | |
| H | 94 | hCoV-19/Scotland/QEUH-54A023/2020\|EPI_ISL_533419\|2 | ···················· | ···················· | ···················· | ···················· | ···················· | ···················· | |
| H | 95 | hCoV-19/England/QEUH-547C8C/2020\|EPI_ISL_533420\|20 | ···················· | ···················· | ···················· | ···················· | ···················· | ···················· | |
| H | 96 | hCoV-19/Scotland/QEUH-548007/2020\|EPI_ISL_533421\|2 | ···················· | ··········A········· | ···················· | ···················· | ···················· | ···················· | |
| H | 97 | hCoV-19/England/QEUH-54D4BB/2020\|EPI_ISL_533422\|20 | ···················· | ···················· | ···················· | ···················· | ···················· | ···················· | |
| H | 98 | hCoV-19/England/QEUH-54DB6B/2020\|EPI_ISL_533424\|20 | ···················· | ···················· | ···················· | ···················· | ···················· | ···················· | |
| H | 99 | hCoV-19/England/QEUH-54AE65/2020\|EPI_ISL_533425\|20 | ···················· | ···················· | ···················· | ···················· | ···················· | ···················· | |
| H | 100 | hCoV-19/England/QEUH-54723F/2020\|EPI_ISL_533426\|20 | ···················· | ···················· | ···················· | ···················· | ···················· | ···················· | |
| H | 101 | hCoV-19/England/QEUH-5470EA/2020\|EPI_ISL_533428\|20 | ···················· | ···················· | ···················· | ···················· | ···················· | ···················· | |
| H | 102 | hCoV-19/England/QEUH-54B9FA/2020\|EPI_ISL_533429\|20 | ···················· | ···················· | ···················· | ···················· | ···················· | ···················· | |
| H | 103 | hCoV-19/England/QEUH-54818C/2020\|EPI_ISL_533430\|20 | ···················· | ···················· | ···················· | ···················· | ···················· | ···················· | |
| H | 104 | hCoV-19/England/QEUH-548B8E/2020\|EPI_ISL_533431\|20 | ···················· | ·A·················· | ···················· | ···················· | ···················· | ···················· | |
| H | 105 | hCoV-19/England/QEUH-548BCA/2020\|EPI_ISL_533433\|20 | ···················· | ···················· | ···················· | ···················· | ···················· | ···················· | |
| H | 106 | hCoV-19/England/203690348/2020\|EPI_ISL_534723\|2020 | ···················· | ···················· | ···················· | ···················· | ···················· | ···················· | |
| H | 107 | hCoV-19/England/203690351/2020\|EPI_ISL_534724\|2020 | ···················· | ···················· | ···················· | ···················· | ···················· | ···················· | |
| H | 108 | hCoV-19/England/203690354/2020\|EPI_ISL_534725\|2020 | ···················· | ···················· | ···················· | ···················· | ···················· | ···················· | |
| H | 109 | hCoV-19/England/203690358/2020\|EPI_ISL_534726\|2020 | ···················· | ···················· | ···················· | ···················· | ···················· | ···················· | |
| H | 110 | hCoV-19/England/203690360/2020\|EPI_ISL_534727\|2020 | ···················· | ···················· | ···················· | ···················· | ···················· | ···················· | |
| H | 111 | hCoV-19/England/203690361/2020\|EPI_ISL_534728\|2020 | ···················· | ···················· | ···················· | ···················· | ···················· | ···················· | |
| H | 112 | hCoV-19/England/203690368/2020\|EPI_ISL_534729\|2020 | ···················· | ···················· | ···················· | ···················· | ···················· | ···················· | |
| H | 113 | hCoV-19/USA/NY-WC-13741-01/2020\|EPI_ISL_534731\|202 | ···················· | ···················· | ···················· | ···················· | ···················· | ···················· | |
| H | 114 | hCoV-19/England/ALDP-98CDFD/2020\|EPI_ISL_534732\|20 | ···················· | ···················· | ···················· | ···················· | ···················· | ···················· | |
| H | 115 | hCoV-19/England/LIVE-1D290B/2020\|EPI_ISL_534748\|20 | ···················· | ···················· | ···················· | ···················· | ···················· | ···················· | |
| H | 116 | hCoV-19/England/LIVE-A97C8/2020\|EPI_ISL_534749\|202 | ···················· | ···················· | ···················· | ···················· | ···················· | ···················· | |
| H | 117 | hCoV-19/England/LIVE-A97D7/2020\|EPI_ISL_534750\|202 | ···················· | ···················· | ···················· | ···················· | ···················· | ···················· | |
| H | 118 | hCoV-19/England/LIVE-A97F5/2020\|EPI_ISL_534751\|202 | ···················· | ·A·················· | ···················· | ···················· | ···················· | ···················· | |
| H | 119 | hCoV-19/England/LIVE-A9801/2020\|EPI_ISL_534752\|202 | ···················· | ·A·················· | ···················· | ···················· | ···················· | ···················· | |
| H | 120 | hCoV-19/England/LIVE-A982F/2020\|EPI_ISL_534753\|202 | ···················· | ···················· | ···················· | ···················· | ···················· | ···················· | |
| H | 121 | hCoV-19/England/LIVE-A983E/2020\|EPI_ISL_534754\|202 | ···················· | ·A·················· | ···················· | ···················· | ···················· | ···················· | |
| H | 122 | hCoV-19/England/LIVE-A984D/2020\|EPI_ISL_534755\|202 | ···················· | ···················· | ···················· | ···················· | ···················· | ···················· | |
| H | 123 | hCoV-19/England/OXON-AF382/2020\|EPI_ISL_534763\|202 | ···················· | ···················· | ···················· | ···················· | ···················· | ···················· | |
| H | 124 | hCoV-19/England/OXON-AF504/2020\|EPI_ISL_534764\|202 | ···················· | ···················· | ···················· | ···················· | ···················· | ···················· | |
| H | 125 | hCoV-19/England/OXON-AF980/2020\|EPI_ISL_534765\|202 | ···················· | ···················· | ···················· | ···················· | ···················· | ···················· | |
| H | 126 | hCoV-19/England/OXON-AFFE8/2020\|EPI_ISL_534766\|202 | ···················· | ···················· | ···················· | ···················· | ···················· | ···················· | |
| H | 127 | hCoV-19/England/OXON-B046D/2020\|EPI_ISL_534767\|202 | ···················· | ···················· | ···················· | ···················· | ···················· | ···················· | |
| H | 128 | hCoV-19/England/OXON-B0764/2020\|EPI_ISL_534768\|202 | ···················· | ···················· | ···················· | ···················· | ···················· | ···················· | |
| H | 129 | hCoV-19/England/OXON-B2762/2020\|EPI_ISL_534769\|202 | ···················· | ···················· | ···················· | ···················· | ···················· | ···················· | |
| H | 130 | hCoV-19/England/OXON-B279F/2020\|EPI_ISL_534770\|202 | ···················· | ···················· | ···················· | ···················· | ···················· | ···················· | |
| H | 131 | hCoV-19/England/OXON-B27CC/2020\|EPI_ISL_534771\|202 | ···················· | ···················· | ···················· | ···················· | ···················· | ···················· | |
| H | 132 | hCoV-19/England/OXON-B27EA/2020\|EPI_ISL_534772\|202 | ···················· | ···················· | ···················· | ···················· | ···················· | ···················· | |
| H | 133 | hCoV-19/England/OXON-B2832/2020\|EPI_ISL_534774\|202 | ···················· | ···················· | ···················· | ···················· | ···················· | ···················· | |
| H | 134 | hCoV-19/England/OXON-B2841/2020\|EPI_ISL_534775\|202 | ···················· | ···················· | ···················· | ···················· | ···················· | ···················· | |
| H | 135 | hCoV-19/England/OXON-B2850/2020\|EPI_ISL_534776\|202 | ···················· | ···················· | ···················· | ···················· | ···················· | ···················· | |
| H | 136 | hCoV-19/England/OXON-B286F/2020\|EPI_ISL_534777\|202 | ···················· | ···················· | ···················· | ···················· | ···················· | ···················· | |
| H | 137 | hCoV-19/England/OXON-B288D/2020\|EPI_ISL_534778\|202 | ···················· | ···················· | ···················· | ···················· | ···················· | ···················· | |
| H | 138 | hCoV-19/England/OXON-B28BA/2020\|EPI_ISL_534779\|202 | ···················· | ···················· | ···················· | ···················· | ···················· | ···················· | |
| H | 139 | hCoV-19/England/OXON-B28E7/2020\|EPI_ISL_534780\|202 | ···················· | ···················· | ···················· | ···················· | ···················· | ···················· | |
| H | 140 | hCoV-19/England/OXON-B294E/2020\|EPI_ISL_534781\|202 | ···················· | ···················· | ···················· | ···················· | ···················· | ···················· | |
| H | 141 | hCoV-19/England/OXON-B298A/2020\|EPI_ISL_534782\|202 | ···················· | ···················· | ···················· | ···················· | ···················· | ···················· | |
| H | 142 | hCoV-19/England/OXON-B2999/2020\|EPI_ISL_534783\|202 | ···················· | ···················· | ···················· | ···················· | ···················· | ···················· | |
| H | 143 | hCoV-19/England/OXON-B29A8/2020\|EPI_ISL_534784\|202 | ···················· | ···················· | ···················· | ···················· | ···················· | ···················· | |
| H | 144 | hCoV-19/England/OXON-B29C6/2020\|EPI_ISL_534785\|202 | ···················· | ···················· | ···················· | ···················· | ···················· | ···················· | |
| H | 145 | hCoV-19/England/OXON-B29E4/2020\|EPI_ISL_534786\|202 | ···················· | ···················· | ···················· | ···················· | ···················· | ···················· | |
| H | 146 | hCoV-19/England/OXON-B2A3C/2020\|EPI_ISL_534787\|202 | ···················· | ···················· | ···················· | ···················· | ···················· | ···················· | |
| H | 147 | hCoV-19/England/OXON-B2A69/2020\|EPI_ISL_534789\|202 | ···················· | ···················· | ···················· | ···················· | ···················· | ···················· | |
| H | 148 | hCoV-19/England/OXON-B2A78/2020\|EPI_ISL_534790\|202 | ···················· | ·············A······ | ···················· | ···················· | ···················· | ···················· | |
| H | 149 | hCoV-19/England/OXON-B2A87/2020\|EPI_ISL_534791\|202 | ···················· | ···················· | ···················· | ···················· | ···················· | ···················· | |
| H | 150 | hCoV-19/England/OXON-B2AC3/2020\|EPI_ISL_534792\|202 | ···················· | ···················· | ···················· | ···················· | ···················· | ···················· | |
| H | 151 | hCoV-19/England/OXON-B2AF0/2020\|EPI_ISL_534793\|202 | ···················· | ···················· | ···················· | ···················· | ···················· | ···················· | |
| H | 152 | hCoV-19/England/OXON-B2B2A/2020\|EPI_ISL_534795\|202 | ···················· | ···················· | ···················· | ···················· | ···················· | ···················· | |
| H | 153 | hCoV-19/England/OXON-B2B48/2020\|EPI_ISL_534796\|202 | ···················· | ···················· | ···················· | ···················· | ···················· | ···················· | |
| H | 154 | hCoV-19/England/OXON-B2B57/2020\|EPI_ISL_534797\|202 | ···················· | ···················· | ···················· | ···················· | ···················· | ···················· | |
| H | 155 | hCoV-19/England/OXON-B2BB1/2020\|EPI_ISL_534798\|202 | ···················· | ···················· | ···················· | ···················· | ···················· | ···················· | |
| H | 156 | hCoV-19/England/OXON-B2BC0/2020\|EPI_ISL_534799\|202 | ···················· | ···················· | ···················· | ···················· | ···················· | ···················· | |
| H | 157 | hCoV-19/England/QEUH-95E28C/2020\|EPI_ISL_537042\|20 | ···················· | ···················· | ···················· | ···················· | ···················· | ···················· | |

| | | | | | | | | |
|---|---|---|---|---|---|---|---|---|
| H | 158 hCoV-19/England/QEUH-95E231/2020\|EPI_ISL_537043\|20 | ···················· | ···················· | ···················· | ···················· | ···················· | ···················· | |
| H | 159 hCoV-19/England/QEUH-95E073/2020\|EPI_ISL_537044\|20 | ···················· | ···················· | ···················· | ···················· | ···················· | ···················· | |
| H | 160 hCoV-19/England/QEUH-95E4D1/2020\|EPI_ISL_537048\|20 | ···················· | ···················· | ···················· | ···················· | ···················· | ···················· | |
| H | 161 hCoV-19/England/QEUH-961514/2020\|EPI_ISL_537049\|20 | ···················· | ···················· | ···················· | ···················· | ···················· | ···················· | |
| H | 162 hCoV-19/England/QEUH-961B03/2020\|EPI_ISL_537050\|20 | ···················· | ···················· | ···················· | ···················· | ···················· | ···················· | |
| H | 163 hCoV-19/England/QEUH-961505/2020\|EPI_ISL_537051\|20 | ···················· | ···················· | ···················· | ···················· | ···················· | ···················· | |
| H | 164 hCoV-19/England/QEUH-95DF2C/2020\|EPI_ISL_537052\|20 | ···················· | ···················· | ···················· | ···················· | ···················· | ···················· | |
| H | 165 hCoV-19/England/QEUH-95DE2F/2020\|EPI_ISL_537053\|20 | ···················· | ···················· | ···················· | ···················· | ···················· | ···················· | |
| H | 166 hCoV-19/England/QEUH-9615E7/2020\|EPI_ISL_537055\|20 | ···················· | ···················· | ···················· | ···················· | ···················· | ···················· | |
| H | 167 hCoV-19/England/QEUH-961DFE/2020\|EPI_ISL_537056\|20 | ···················· | ·A·················· | ···················· | ···················· | ···················· | ···················· | |
| H | 168 hCoV-19/England/QEUH-9614BD/2020\|EPI_ISL_537057\|20 | ···················· | ·A·················· | ···················· | ···················· | ···················· | ···················· | |
| H | 169 hCoV-19/England/QEUH-95DEF2/2020\|EPI_ISL_537058\|20 | ···················· | ···················· | ···················· | ···················· | ···················· | ···················· | |
| H | 170 hCoV-19/England/QEUH-961D2B/2020\|EPI_ISL_537059\|20 | ···················· | ···················· | ···················· | ···················· | ···················· | ···················· | |
| H | 171 hCoV-19/England/QEUH-961857/2020\|EPI_ISL_537060\|20 | ···················· | ···················· | ···················· | ···················· | ···················· | ···················· | |
| H | 172 hCoV-19/England/QEUH-961D3A/2020\|EPI_ISL_537062\|20 | ···················· | ···················· | ··········T········· | ···················· | ···················· | ···················· | |
| H | 173 hCoV-19/England/QEUH-9618EE/2020\|EPI_ISL_537064\|20 | ···················· | ···················· | ···················· | ···················· | ···················· | ···················· | |
| H | 174 hCoV-19/Northern Ireland/QEUH-961ABB/2020\|EPI_ISL_ | ···················· | ···················· | ···················· | ···················· | ···················· | ···················· | |
| H | 175 hCoV-19/Northern Ireland/QEUH-961BE5/2020\|EPI_ISL_ | ···················· | ···················· | ···················· | ···················· | ···················· | ···················· | |
| H | 176 hCoV-19/England/QEUH-95DF3B/2020\|EPI_ISL_537067\|20 | ···················· | ···················· | ···················· | ···················· | ···················· | ···················· | |
| H | 177 hCoV-19/England/QEUH-95DCCB/2020\|EPI_ISL_537068\|20 | ···················· | ···················· | ···················· | ···················· | ···················· | ···················· | |
| H | 178 hCoV-19/England/QEUH-96170F/2020\|EPI_ISL_537069\|20 | ···················· | ···················· | ···················· | ···················· | ···················· | ···················· | |
| H | 179 hCoV-19/England/QEUH-95E4E0/2020\|EPI_ISL_537071\|20 | ···················· | ···················· | ···················· | ···················· | ···················· | ···················· | |
| H | 180 hCoV-19/Canada/ON-OICR-SLB2851/2020\|EPI_ISL_538370 | ···················· | ···················· | ···················· | ···················· | ···················· | ···················· | |
| H | 181 hCoV-19/Canada/ON-OICR-SLB2852/2020\|EPI_ISL_538371 | ···················· | ···················· | ···················· | ···················· | ···················· | ···················· | |
| H | 182 hCoV-19/Canada/ON-OICR-SLB2853/2020\|EPI_ISL_538372 | ···················· | ···················· | ···················· | ···················· | ···················· | ···················· | |
| H | 183 hCoV-19/Canada/ON-OICR-SLB2854/2020\|EPI_ISL_538373 | ···················· | ···················· | ···················· | ···················· | ···················· | ···················· | |
| H | 184 hCoV-19/Canada/ON-OICR-SLB2855/2020\|EPI_ISL_538374 | ···················· | ···················· | ···················· | ···················· | ···················· | ···················· | |
| H | 185 hCoV-19/Canada/ON-OICR-SLB2856/2020\|EPI_ISL_538375 | ···················· | ···················· | ···················· | ···················· | ···················· | ···················· | |
| H | 186 hCoV-19/Canada/ON-OICR-SLB2857/2020\|EPI_ISL_538376 | ···················· | ···················· | ···················· | ···················· | ···················· | ···················· | |
| H | 187 hCoV-19/Canada/ON-OICR-SLB2858/2020\|EPI_ISL_538377 | ···················· | ···················· | ···················· | ···················· | ···················· | ···················· | |
| H | 188 hCoV-19/Canada/ON-OICR-SLB2859/2020\|EPI_ISL_538378 | ···················· | ···················· | ···················· | ···················· | ···················· | ···················· | |
| H | 189 hCoV-19/Canada/ON-OICR-SLB2860/2020\|EPI_ISL_538379 | ···················· | ···················· | ···················· | ···················· | ···················· | ···················· | |
| H | 190 hCoV-19/Canada/ON-OICR-SLB2863/2020\|EPI_ISL_538380 | ···················· | ···················· | ···················· | ···················· | ···················· | ···················· | |
| H | 191 hCoV-19/Canada/ON-OICR-SLB2864/2020\|EPI_ISL_538381 | ···················· | ···················· | ···················· | ···················· | ···················· | ···················· | |
| H | 192 hCoV-19/Canada/ON-OICR-SLB2865/2020\|EPI_ISL_538382 | ···················· | ···················· | ···················· | ···················· | ···················· | ···················· | |
| H | 193 hCoV-19/Canada/ON-OICR-SLB2866/2020\|EPI_ISL_538383 | ···················· | ···················· | ···················· | ···················· | ···················· | ···················· | |
| H | 194 hCoV-19/Canada/ON-OICR-SLB2867/2020\|EPI_ISL_538384 | ···················· | ···················· | ···················· | ···················· | ···················· | ···················· | |
| H | 195 hCoV-19/Canada/ON-OICR-SLB2868/2020\|EPI_ISL_538385 | ···················· | ···················· | ···················· | ···················· | ···················· | ···················· | |
| H | 196 hCoV-19/Canada/ON-OICR-SLB2869/2020\|EPI_ISL_538386 | ···················· | ···················· | ···················· | ···················· | ···················· | ···················· | |
| H | 197 hCoV-19/USA/SC-DHEC-0766/2020\|EPI_ISL_538390\|2020- | ···················· | ···················· | ···················· | ···················· | ···················· | ···················· | |
| H | 198 hCoV-19/USA/SC-DHEC-0768/2020\|EPI_ISL_538391\|2020- | ···················· | ···················· | ···················· | ···················· | ···················· | ···················· | |
| H | 199 hCoV-19/USA/SC-DHEC-0772/2020\|EPI_ISL_538392\|2020- | ···················· | ···················· | ···················· | ···················· | ···················· | ···················· | |
| H | 200 hCoV-19/USA/SC-DHEC-0774/2020\|EPI_ISL_538393\|2020- | ···················· | ···················· | ···················· | ···················· | ···················· | ···················· | |
| H | 201 hCoV-19/USA/SC-DHEC-0776/2020\|EPI_ISL_538395\|2020- | ···················· | ···················· | ···················· | ···················· | ···················· | ···················· | |
| H | 202 hCoV-19/USA/SC-DHEC-0777/2020\|EPI_ISL_538396\|2020- | ···················· | ···················· | ···················· | ···················· | ···················· | ···················· | |
| H | 203 hCoV-19/USA/SC-DHEC-0781/2020\|EPI_ISL_538397\|2020- | ···················· | ···················· | ···················· | ···················· | ···················· | ···················· | |
| H | 204 hCoV-19/USA/SC-DHEC-0863/2020\|EPI_ISL_538399\|2020- | ···················· | ·A·················· | ···················· | ···················· | ···················· | ···················· | |
| H | 205 hCoV-19/Germany/NW-HHU-84/2020\|EPI_ISL_539600\|2020 | ···················· | ···················· | ···················· | ···················· | ···················· | ···················· | |
| H | 206 hCoV-19/Germany/NW-HHU-85/2020\|EPI_ISL_539601\|2020 | ···················· | ···················· | ···················· | ···················· | ···················· | ···················· | |
| H | 207 hCoV-19/Germany/NW-HHU-86/2020\|EPI_ISL_539602\|2020 | ···················· | ···················· | ···················· | ···················· | ···················· | ···················· | |
| H | 208 hCoV-19/Germany/NW-HHU-87/2020\|EPI_ISL_539603\|2020 | ···················· | ···················· | ···················· | ···················· | ···················· | ···················· | |
| H | 209 hCoV-19/Germany/NW-HHU-88/2020\|EPI_ISL_539604\|2020 | ···················· | ···················· | ···················· | ···················· | ···················· | ···················· | |
| H | 210 hCoV-19/Germany/NW-HHU-89/2020\|EPI_ISL_539605\|2020 | ···················· | ···················· | ···················· | ···················· | ···················· | ···················· | |
| H | 211 hCoV-19/Germany/NW-HHU-90/2020\|EPI_ISL_539606\|2020 | ···················· | ···················· | ···················· | ···················· | ···················· | ···················· | |
| H | 212 hCoV-19/Germany/NW-HHU-91/2020\|EPI_ISL_539607\|2020 | ···················· | ···················· | ···················· | ···················· | ···················· | ···················· | |
| H | 213 hCoV-19/Germany/NW-HHU-93/2020\|EPI_ISL_539609\|2020 | ···················· | ···················· | ···················· | ···················· | ···················· | ···················· | |
| H | 214 hCoV-19/Germany/NW-HHU-94/2020\|EPI_ISL_539610\|2020 | ···················· | ···················· | ···················· | ···················· | ···················· | ···················· | |
| H | 215 hCoV-19/Germany/NW-HHU-95/2020\|EPI_ISL_539611\|2020 | ···················· | ···················· | ···················· | ···················· | ···················· | ···················· | |
| H | 216 hCoV-19/Germany/NW-HHU-97/2020\|EPI_ISL_539613\|2020 | ···················· | ···················· | ···················· | ···················· | ···················· | ···················· | |
| H | 217 hCoV-19/Germany/NW-HHU-98/2020\|EPI_ISL_539614\|2020 | ···················· | ···················· | ···················· | ···················· | ···················· | ···················· | |
| H | 218 hCoV-19/Germany/NW-HHU-99/2020\|EPI_ISL_539615\|2020 | ···················· | ···················· | ···················· | ···················· | ···················· | ···················· | |
| H | 219 hCoV-19/India/TG-CCMB-AA147/2020\|EPI_ISL_539616\|20 | ···················· | ···················· | ···················· | ···················· | ···················· | ···················· | |
| H | 220 hCoV-19/India/TG-CCMB-AB465/2020\|EPI_ISL_539617\|20 | ···················· | ···················· | ···················· | ···················· | ···················· | ···················· | |
| H | 221 hCoV-19/England/MILK-9924A4/2020\|EPI_ISL_540685\|20 | ···················· | ···················· | ···················· | ···················· | ···················· | ···················· | |
| H | 222 hCoV-19/England/MILK-99250B/2020\|EPI_ISL_540686\|20 | ···················· | ···················· | ···················· | ···················· | ···················· | ···················· | |
| H | 223 hCoV-19/England/MILK-99251A/2020\|EPI_ISL_540687\|20 | ···················· | ·A·················· | ···················· | ···················· | ···················· | ··A················· | |
| H | 224 hCoV-19/England/MILK-992529/2020\|EPI_ISL_540688\|20 | ···················· | ···················· | ···················· | ···················· | ···················· | ···················· | |
| H | 225 hCoV-19/England/MILK-992538/2020\|EPI_ISL_540689\|20 | ···················· | ···················· | ···················· | ···················· | ···················· | ···················· | |
| H | 227 hCoV-19/England/NORW-EEA55/2020\|EPI_ISL_540692\|202 | ···················· | ···················· | ···················· | ···················· | ···················· | ···················· | |
| H | 228 hCoV-19/England/NORW-EEA82/2020\|EPI_ISL_540693\|202 | ···················· | ···················· | ···················· | ···················· | ···················· | ···················· | |
| H | 229 hCoV-19/England/NORW-EEC40/2020\|EPI_ISL_540694\|202 | ···················· | ···················· | ···················· | ···················· | NNN················· | ···················· | 1 |
| H | 230 hCoV-19/England/NORW-EEC7D/2020\|EPI_ISL_540695\|202 | ···················· | ···················· | ···················· | ···················· | ···················· | ···················· | |
| H | 234 hCoV-19/Spain/MD-IBV-99007129/2020\|EPI_ISL_541903\| | ···················· | ···················· | ···················· | ···················· | ···················· | ···················· | |
| H | 235 hCoV-19/Spain/MD-IBV-99007131/2020\|EPI_ISL_541904\| | ···················· | ···················· | ···················· | ···················· | ···················· | ···················· | |
| H | 244 hCoV-19/Spain/MD-IBV-99007144/2020\|EPI_ISL_541916\| | ···················· | ···················· | ···················· | ···················· | ···················· | ···················· | |
| H | 254 hCoV-19/Spain/MD-IBV-99007157/2020\|EPI_ISL_541927\| | ···················· | ···················· | ···················· | ···················· | ···················· | ···················· | |
| H | 257 hCoV-19/England/OXON-B2C09/2020\|EPI_ISL_534800\|202 | ···················· | ···················· | ···················· | ···················· | ···················· | ···················· | |
| H | 258 hCoV-19/England/OXON-B2C18/2020\|EPI_ISL_534801\|202 | ···················· | ···················· | ···················· | ···················· | ···················· | ···················· | |
| H | 259 hCoV-19/England/OXON-B2C63/2020\|EPI_ISL_534802\|202 | ···················· | ···················· | ···················· | ···················· | ···················· | ···················· | |
| H | 260 hCoV-19/England/OXON-B2C90/2020\|EPI_ISL_534803\|202 | ···················· | ···················· | ···················· | ···················· | ···················· | ···················· | |

| | | | | | | | | |
|---|---|---|---|---|---|---|---|---|
| H | 261 | hCoV-19/England/OXON-B2CAF/2020\|EPI_ISL_534804\|202 | .................... | .................... | ..................... | .................... | .................... | .................... |
| H | 262 | hCoV-19/England/OXON-B2CBE/2020\|EPI_ISL_534805\|202 | .................... | .................... | ..................... | .................... | .................... | .................... |
| H | 263 | hCoV-19/England/OXON-B2CCD/2020\|EPI_ISL_534806\|202 | .................... | .................... | ..................... | .................... | .................... | .................... |
| H | 264 | hCoV-19/England/OXON-B2CDC/2020\|EPI_ISL_534807\|202 | .................... | .................... | ..................... | .................... | .................... | .................... |
| H | 265 | hCoV-19/England/OXON-B2D06/2020\|EPI_ISL_534808\|202 | .................... | .................... | ..................... | .................... | .................... | .................... |
| H | 266 | hCoV-19/England/OXON-B2D15/2020\|EPI_ISL_534809\|202 | .................... | .................... | ..................... | .................... | .................... | .................... |
| H | 267 | hCoV-19/England/OXON-B2D24/2020\|EPI_ISL_534810\|202 | .................... | .................... | ..................... | .................... | .................... | .................... |
| H | 268 | hCoV-19/England/OXON-B2D33/2020\|EPI_ISL_534811\|202 | .................... | .................... | ..................... | .................... | .................... | .................... |
| H | 269 | hCoV-19/England/OXON-B2D7F/2020\|EPI_ISL_534813\|202 | .................... | .................... | ..................... | .................... | .................... | .................... |
| H | 270 | hCoV-19/England/OXON-B2D9D/2020\|EPI_ISL_534814\|202 | .................... | .................... | ..................... | .................... | .................... | .................... |
| H | 271 | hCoV-19/England/OXON-B2DD9/2020\|EPI_ISL_534815\|202 | .................... | .................... | ..................... | .................... | .................... | .................... |
| H | 272 | hCoV-19/England/OXON-B2DF7/2020\|EPI_ISL_534816\|202 | .................... | .................... | ..................... | .................... | .................... | .................... |
| H | 273 | hCoV-19/England/OXON-B2E12/2020\|EPI_ISL_534817\|202 | .................... | .................... | ..................... | .................... | .................... | .................... |
| H | 274 | hCoV-19/England/OXON-B2E6D/2020\|EPI_ISL_534819\|202 | .................... | .................... | ..................... | .................... | .................... | .................... |
| H | 275 | hCoV-19/England/OXON-B2E7C/2020\|EPI_ISL_534820\|202 | .................... | .................... | ..................... | .................... | .................... | .................... |
| H | 276 | hCoV-19/England/OXON-B2E8B/2020\|EPI_ISL_534821\|202 | .................... | .................... | ..................... | .................... | .................... | .................... |
| H | 277 | hCoV-19/England/OXON-B2EB8/2020\|EPI_ISL_534822\|202 | .................... | ..........A......... | ..................... | .................... | .................... | .................... |
| H | 278 | hCoV-19/England/OXON-B2EC7/2020\|EPI_ISL_534823\|202 | .................... | .................... | ..................... | .................... | .................... | .................... |
| H | 279 | hCoV-19/England/OXON-B2F00/2020\|EPI_ISL_534825\|202 | .................... | .................... | ..................... | .................... | .................... | .................... |
| H | 280 | hCoV-19/England/OXON-B2F1F/2020\|EPI_ISL_534826\|202 | .................... | .................... | ..................... | .................... | .................... | .................... |
| H | 281 | hCoV-19/England/OXON-B2F2E/2020\|EPI_ISL_534827\|202 | .................... | .A.................. | ..................... | .................... | .................... | .................... |
| H | 282 | hCoV-19/England/OXON-B2F4C/2020\|EPI_ISL_534828\|202 | .................... | .................... | ..................... | .................... | .................... | .................... |
| H | 283 | hCoV-19/England/OXON-B2F5B/2020\|EPI_ISL_534829\|202 | .................... | .................... | ..................... | .................... | .................... | .................... |
| H | 284 | hCoV-19/England/OXON-B2F79/2020\|EPI_ISL_534830\|202 | .................... | .................... | ..................... | .................... | .................... | .................... |
| H | 285 | hCoV-19/England/OXON-B2F88/2020\|EPI_ISL_534831\|202 | .................... | .................... | ..................... | .................... | .................... | .................... |
| H | 286 | hCoV-19/England/OXON-B2FE2/2020\|EPI_ISL_534833\|202 | .................... | .................... | ..................... | .................... | .................... | .................... |
| H | 287 | hCoV-19/England/OXON-B3048/2020\|EPI_ISL_534834\|202 | .................... | .................... | ..................... | .................... | .................... | .................... |
| H | 288 | hCoV-19/England/OXON-B3075/2020\|EPI_ISL_534835\|202 | .................... | .................... | ..................... | .................... | .................... | .................... |
| H | 289 | hCoV-19/England/OXON-B3084/2020\|EPI_ISL_534836\|202 | .................... | .................... | ..................... | .................... | .................... | .................... |
| H | 290 | hCoV-19/England/OXON-B30DF/2020\|EPI_ISL_534837\|202 | .................... | .................... | ..................... | .................... | .................... | .................... |
| H | 291 | hCoV-19/England/OXON-B3118/2020\|EPI_ISL_534838\|202 | .................... | .................... | ..................... | .................... | .................... | .................... |
| H | 292 | hCoV-19/England/OXON-B3145/2020\|EPI_ISL_534839\|202 | .................... | .................... | ..................... | .................... | .................... | .................... |
| H | 293 | hCoV-19/England/OXON-B31AF/2020\|EPI_ISL_534840\|202 | .................... | .................... | ..................... | .................... | .................... | .................... |
| H | 294 | hCoV-19/England/OXON-B31BE/2020\|EPI_ISL_534841\|202 | .................... | .................... | ..................... | .................... | .................... | .................... |
| H | 295 | hCoV-19/England/OXON-B31DC/2020\|EPI_ISL_534842\|202 | .................... | .................... | ..................... | .................... | .................... | .................... |
| H | 296 | hCoV-19/England/OXON-B31EB/2020\|EPI_ISL_534843\|202 | .................... | .................... | ..................... | .................... | .................... | .................... |
| H | 297 | hCoV-19/England/QEUH-95DFD1/2020\|EPI_ISL_537000\|20 | .................... | .................... | ..................... | .................... | .................... | .................... |
| H | 298 | hCoV-19/England/QEUH-95E398/2020\|EPI_ISL_537001\|20 | .................... | .A.................. | ..................... | .................... | .................... | ...A................ |
| H | 299 | hCoV-19/England/QEUH-961B9A/2020\|EPI_ISL_537002\|20 | .................... | .................... | ..................... | .................... | .................... | .................... |
| H | 300 | hCoV-19/England/QEUH-95E116/2020\|EPI_ISL_537003\|20 | .................... | .................... | ..................... | .................... | .................... | .................... |
| H | 301 | hCoV-19/England/QEUH-95E3C5/2020\|EPI_ISL_537004\|20 | .................... | .................... | ..................... | .................... | .................... | .................... |
| H | 302 | hCoV-19/England/QEUH-95E3E3/2020\|EPI_ISL_537005\|20 | .................... | .................... | ..................... | .................... | .................... | .................... |
| H | 303 | hCoV-19/England/QEUH-95E0DD/2020\|EPI_ISL_537006\|20 | .................... | .................... | ..................... | .................... | .................... | .................... |
| H | 304 | hCoV-19/England/QEUH-961DD0/2020\|EPI_ISL_537007\|20 | .................... | .................... | ..................... | .................... | .................... | .................... |
| H | 305 | hCoV-19/England/QEUH-95DF68/2020\|EPI_ISL_537008\|20 | .................... | .................... | ..................... | .................... | .................... | .................... |
| H | 306 | hCoV-19/England/QEUH-961778/2020\|EPI_ISL_537009\|20 | .................... | .................... | ..................... | .................... | .................... | .................... |
| H | 307 | hCoV-19/England/QEUH-95E529/2020\|EPI_ISL_537011\|20 | .................... | .................... | ..................... | .................... | .................... | .................... |
| H | 308 | hCoV-19/England/QEUH-961A06/2020\|EPI_ISL_537012\|20 | .................... | .................... | ..................... | .................... | .................... | .................... |
| H | 309 | hCoV-19/England/QEUH-961796/2020\|EPI_ISL_537013\|20 | .................... | .................... | ..................... | .................... | .................... | .................... |
| H | 310 | hCoV-19/England/QEUH-95E41D/2020\|EPI_ISL_537014\|20 | .................... | .................... | ..................... | .................... | .................... | .................... |
| H | 311 | hCoV-19/England/QEUH-95DFE0/2020\|EPI_ISL_537015\|20 | .................... | .................... | ..................... | .................... | .................... | .................... |
| H | 312 | hCoV-19/Wales/QEUH-96163F/2020\|EPI_ISL_537016\|2020 | .................... | .................... | ..................... | .................... | .................... | .................... |
| H | 313 | hCoV-19/England/QEUH-9614EA/2020\|EPI_ISL_537018\|20 | .................... | .................... | ..................... | .................... | .................... | .................... |
| H | 314 | hCoV-19/England/QEUH-95DD9B/2020\|EPI_ISL_537019\|20 | .................... | .................... | ..................... | .................... | .................... | .................... |
| H | 315 | hCoV-19/England/QEUH-95DF77/2020\|EPI_ISL_537021\|20 | .................... | .................... | ..................... | .................... | .................... | .................... |
| H | 316 | hCoV-19/England/QEUH-95E046/2020\|EPI_ISL_537022\|20 | .................... | .................... | ..................... | .................... | .................... | .................... |
| H | 317 | hCoV-19/Northern Ireland/QEUH-961D58/2020\|EPI_ISL_ | .................... | .................... | ..................... | .................... | .................... | .................... |
| H | 318 | hCoV-19/England/QEUH-961954/2020\|EPI_ISL_537025\|20 | .................... | .................... | ..................... | .................... | .................... | .................... |
| H | 319 | hCoV-19/England/QEUH-9616A8/2020\|EPI_ISL_537026\|20 | .................... | .................... | ..................... | .................... | .................... | .................... |
| H | 320 | hCoV-19/England/QEUH-961462/2020\|EPI_ISL_537027\|20 | .................... | .................... | ..................... | .................... | .................... | .................... |
| H | 321 | hCoV-19/Northern Ireland/QEUH-961C97/2020\|EPI_ISL_ | .................... | .................... | ..................... | .................... | .................... | .................... |
| H | 322 | hCoV-19/England/QEUH-961963/2020\|EPI_ISL_537029\|20 | .................... | .................... | ..................... | .................... | .................... | .................... |
| H | 323 | hCoV-19/England/QEUH-961D1C/2020\|EPI_ISL_537070\|20 | .................... | .................... | ..................... | .................... | .................... | .................... |
| H | 324 | hCoV-19/England/QEUH-961AE8/2020\|EPI_ISL_537072\|20 | .................... | .................... | ..................... | .................... | .................... | .................... |
| H | 325 | hCoV-19/England/QEUH-95DD7D/2020\|EPI_ISL_537073\|20 | .................... | .A.................. | ..................... | .................... | .................... | .................... |
| H | 326 | hCoV-19/England/QEUH-9619FA/2020\|EPI_ISL_537075\|20 | .................... | .................... | ..................... | .................... | .................... | .................... |
| H | 327 | hCoV-19/England/QEUH-95DDE6/2020\|EPI_ISL_537076\|20 | .................... | .................... | ..................... | .................... | .................... | .................... |
| H | 328 | hCoV-19/England/QEUH-961699/2020\|EPI_ISL_537078\|20 | .................... | .................... | ..................... | .................... | .................... | .................... |
| H | 329 | hCoV-19/England/QEUH-9617C3/2020\|EPI_ISL_537079\|20 | .................... | .................... | ..................... | .................... | .................... | .................... |
| H | 330 | hCoV-19/England/QEUH-961B21/2020\|EPI_ISL_537080\|20 | .................... | .................... | ..................... | .................... | .................... | .................... |
| H | 331 | hCoV-19/England/QEUH-961E73/2020\|EPI_ISL_537081\|20 | .................... | .................... | ..................... | .................... | .................... | .................... |
| H | 332 | hCoV-19/England/QEUH-95DED4/2020\|EPI_ISL_537082\|20 | .................... | .................... | ..................... | .................... | .................... | .................... |
| H | 333 | hCoV-19/England/QEUH-961CF1/2020\|EPI_ISL_537083\|20 | .................... | .................... | ..................... | .................... | .................... | .................... |
| H | 334 | hCoV-19/England/QEUH-9619DC/2020\|EPI_ISL_537084\|20 | .................... | .................... | ..................... | .................... | .................... | .................... |
| H | 335 | hCoV-19/England/QEUH-96166C/2020\|EPI_ISL_537085\|20 | .................... | .................... | ..................... | .................... | .................... | .................... |
| H | 336 | hCoV-19/England/QEUH-95DE98/2020\|EPI_ISL_537086\|20 | .................... | .................... | ..................... | .................... | .................... | .................... |
| H | 337 | hCoV-19/England/QEUH-95DEE3/2020\|EPI_ISL_537087\|20 | .................... | .................... | ..................... | .................... | .................... | .................... |
| H | 338 | hCoV-19/England/QEUH-95DE89/2020\|EPI_ISL_537088\|20 | .................... | .................... | ..................... | .................... | .................... | .................... |
| H | 339 | hCoV-19/England/QEUH-96159C/2020\|EPI_ISL_537089\|20 | .................... | .................... | ..................... | .................... | .................... | .................... |
| H | 340 | hCoV-19/England/QEUH-95DD31/2020\|EPI_ISL_537090\|20 | .................... | .A.................. | ..................... | .................... | .................... | .................... |

| | | | | | | | | |
|---|---|---|---|---|---|---|---|---|
| H | 341 | hCoV-19/England/QEUH-95E2C8/2020\|EPI_ISL_537091\|20 | .................... | .................... | .................... | .................... | .................... | .................... |
| H | 342 | hCoV-19/England/QEUH-961541/2020\|EPI_ISL_537092\|20 | .................... | .................... | .................... | .................... | .................... | .................... |
| H | 343 | hCoV-19/Wales/QEUH-961E55/2020\|EPI_ISL_537093\|2020 | .................... | .................... | .................... | .................... | .................... | .................... |
| H | 344 | hCoV-19/Northern Ireland/QEUH-961ACA/2020\|EPI_ISL_ | .................... | .................... | .................... | .................... | .................... | .................... |
| H | 345 | hCoV-19/England/QEUH-95DD04/2020\|EPI_ISL_537095\|20 | .................... | .................... | .................... | .................... | .................... | .................... |
| H | 346 | hCoV-19/Northern Ireland/QEUH-961A42/2020\|EPI_ISL_ | .................... | .................... | .................... | .................... | .................... | .................... |
| H | 347 | hCoV-19/England/QEUH-961945/2020\|EPI_ISL_537097\|20 | .................... | .................... | .................... | .................... | .................... | .................... |
| H | 348 | hCoV-19/England/QEUH-95E538/2020\|EPI_ISL_537098\|20 | .................... | .................... | .................... | .................... | .................... | .................... |
| H | 349 | hCoV-19/England/QEUH-961C1F/2020\|EPI_ISL_537099\|20 | .................... | .................... | .................... | .................... | .................... | .................... |
| H | 350 | hCoV-19/USA/TX-DSHS-0371/2020\|EPI_ISL_538307\|2020- | .................... | .................... | .................... | .................... | .................... | .................... |
| H | 351 | hCoV-19/Canada/ON-OICR-SLB2787/2020\|EPI_ISL_538321 | .................... | .................... | .................... | .................... | .................... | .................... |
| H | 352 | hCoV-19/Canada/ON-OICR-SLB2788/2020\|EPI_ISL_538322 | .................... | .................... | .................... | .................... | .................... | .................... |
| H | 353 | hCoV-19/Canada/ON-OICR-SLB2789/2020\|EPI_ISL_538323 | .................... | .................... | .................... | .................... | .................... | .................... |
| H | 354 | hCoV-19/Canada/ON-OICR-SLB2790/2020\|EPI_ISL_538324 | .................... | .................... | .................... | .................... | .................... | .................... |
| H | 355 | hCoV-19/Canada/ON-OICR-SLB2791/2020\|EPI_ISL_538325 | .................... | .................... | .................... | .................... | .................... | .................... |
| H | 356 | hCoV-19/Canada/ON-OICR-SLB2792/2020\|EPI_ISL_538326 | .................... | .................... | .................... | .................... | .................... | .................... |
| H | 357 | hCoV-19/Canada/ON-OICR-SLB2793/2020\|EPI_ISL_538327 | .................... | .................... | .................... | .................... | .................... | .................... |
| H | 358 | hCoV-19/Canada/ON-OICR-SLB2796/2020\|EPI_ISL_538328 | .................... | .................... | .................... | .................... | .................... | .................... |
| H | 359 | hCoV-19/Canada/ON-OICR-SLB2797/2020\|EPI_ISL_538329 | .................... | .................... | .................... | .................... | .................... | .................... |
| H | 360 | hCoV-19/Canada/ON-OICR-SLB2798/2020\|EPI_ISL_538330 | .................... | .................... | .................... | .................... | .................... | .................... |
| H | 361 | hCoV-19/Canada/ON-OICR-SLB2799/2020\|EPI_ISL_538331 | .................... | .................... | .................... | .................... | .................... | .................... |
| H | 362 | hCoV-19/Canada/ON-OICR-SLB2800/2020\|EPI_ISL_538332 | .................... | .................... | .................... | .................... | .................... | .................... |
| H | 363 | hCoV-19/Canada/ON-OICR-SLB2802/2020\|EPI_ISL_538333 | .................... | .................... | .................... | .................... | .................... | .................... |
| H | 364 | hCoV-19/Canada/ON-OICR-SLB2804/2020\|EPI_ISL_538335 | .................... | .................... | .................... | .................... | .................... | .................... |
| H | 365 | hCoV-19/Canada/ON-OICR-SLB2806/2020\|EPI_ISL_538336 | .................... | .................... | .................... | .................... | .................... | .................... |
| H | 366 | hCoV-19/Canada/ON-OICR-SLB2809/2020\|EPI_ISL_538338 | .................... | .................... | .................... | .................... | .................... | .................... |
| H | 367 | hCoV-19/Canada/ON-OICR-SLB2811/2020\|EPI_ISL_538339 | .................... | .................... | .................... | .................... | .................... | .................... |
| H | 368 | hCoV-19/Canada/ON-OICR-SLB2812/2020\|EPI_ISL_538340 | .................... | .................... | .................... | .................... | .................... | .................... |
| H | 369 | hCoV-19/Canada/ON-OICR-SLB2813/2020\|EPI_ISL_538341 | .................... | .................... | .................... | .................... | .................... | .................... |
| H | 370 | hCoV-19/Canada/ON-OICR-SLB2814/2020\|EPI_ISL_538342 | .................... | .................... | .................... | .................... | .................... | .................... |
| H | 371 | hCoV-19/Canada/ON-OICR-SLB2815/2020\|EPI_ISL_538343 | .................... | .................... | .................... | .................... | .................... | .................... |
| H | 372 | hCoV-19/Canada/ON-OICR-SLB2817/2020\|EPI_ISL_538345 | .................... | .................... | .................... | .................... | .................... | .................... |
| H | 373 | hCoV-19/Canada/ON-OICR-SLB2818/2020\|EPI_ISL_538346 | .................... | .................... | .................... | .................... | .................... | .................... |
| H | 374 | hCoV-19/Canada/ON-OICR-SLB2819/2020\|EPI_ISL_538347 | .................... | .................... | .................... | .................... | .................... | .................... |
| H | 375 | hCoV-19/Canada/ON-OICR-SLB2820/2020\|EPI_ISL_538348 | .................... | .................... | .................... | .................... | .................... | .................... |
| H | 376 | hCoV-19/Canada/ON-OICR-SLB2821/2020\|EPI_ISL_538349 | .................... | .................... | .................... | .................... | .................... | .................... |
| H | 377 | hCoV-19/Canada/ON-OICR-SLB2822/2020\|EPI_ISL_538350 | .................... | .................... | .................... | .................... | .................... | .................... |
| H | 378 | hCoV-19/Canada/ON-OICR-SLB2823/2020\|EPI_ISL_538351 | .................... | .................... | .................... | .................... | .................... | .................... |
| H | 379 | hCoV-19/Canada/ON-OICR-SLB2827/2020\|EPI_ISL_538353 | .................... | .................... | .................... | .................... | .................... | .................... |
| H | 380 | hCoV-19/Canada/ON-OICR-SLB2829/2020\|EPI_ISL_538355 | .................... | .................... | .................... | .................... | .................... | .................... |
| H | 381 | hCoV-19/Canada/ON-OICR-SLB2830/2020\|EPI_ISL_538356 | .................... | .................... | .................... | .................... | .................... | .................... |
| H | 382 | hCoV-19/Canada/ON-OICR-SLB2832/2020\|EPI_ISL_538358 | .................... | .................... | .................... | .................... | .................... | .................... |
| H | 383 | hCoV-19/Canada/ON-OICR-SLB2835/2020\|EPI_ISL_538359 | .................... | .................... | .................... | .................... | .................... | .................... |
| H | 384 | hCoV-19/India/TG-CCMB-AB583/2020\|EPI_ISL_539618\|20 | .................... | .................... | .................... | .................... | .................... | .................... |
| H | 385 | hCoV-19/India/TG-CCMB-AC539/2020\|EPI_ISL_539619\|20 | .................... | .................... | .................... | .................... | .................... | .................... |
| H | 386 | hCoV-19/India/TG-CCMB-R249/2020\|EPI_ISL_539620\|202 | .................... | .................... | .................... | .................... | .................... | .................... |
| H | 387 | hCoV-19/India/TG-CCMB-R220/2020\|EPI_ISL_539621\|202 | .................... | .................... | .................... | .................... | .................... | .................... |
| H | 388 | hCoV-19/India/TG-CCMB-R159/2020\|EPI_ISL_539622\|202 | .................... | .................... | .................... | .................... | .................... | .................... |
| H | 389 | hCoV-19/India/TG-CCMB-R178/2020\|EPI_ISL_539623\|202 | .................... | .................... | .................... | .................... | .................... | .................... |
| H | 390 | hCoV-19/India/TG-CCMB-R208/2020\|EPI_ISL_539624\|202 | .................... | .................... | .................... | .................... | .................... | .................... |
| H | 391 | hCoV-19/India/TG-CCMB-R91/2020\|EPI_ISL_539625\|2020 | .................... | .................... | .................... | .................... | .................... | .................... |
| H | 392 | hCoV-19/India/TG-CCMB-R320/2020\|EPI_ISL_539626\|202 | .................... | .................... | .................... | .................... | .................... | .................... |
| H | 393 | hCoV-19/India/TG-CCMB-AB443/2020\|EPI_ISL_539627\|20 | .................... | .................... | .................... | .................... | .................... | .................... |
| H | 394 | hCoV-19/India/TG-CCMB-AA527/2020\|EPI_ISL_539628\|20 | .................... | .................... | .................... | .................... | .................... | .................... |
| H | 395 | hCoV-19/India/TG-CCMB-AB520/2020\|EPI_ISL_539629\|20 | .................... | .................... | .................... | .................... | .................... | .................... |
| H | 396 | hCoV-19/India/TG-CCMB-R394/2020\|EPI_ISL_539630\|202 | .................... | .................... | .................... | .................... | .................... | .................... |
| H | 397 | hCoV-19/India/TG-CCMB-AB364/2020\|EPI_ISL_539631\|20 | .................... | .................... | .................... | .................... | .................... | .................... |
| H | 398 | hCoV-19/India/TG-CCMB-AB352/2020\|EPI_ISL_539632\|20 | .................... | .................... | .................... | .................... | .................... | .................... |
| H | 399 | hCoV-19/India/TG-CCMB-AA560/2020\|EPI_ISL_539633\|20 | .................... | .................... | .................... | .................... | .................... | .................... |
| H | 400 | hCoV-19/India/TG-CCMB-AA562/2020\|EPI_ISL_539634\|20 | .................... | .................... | .................... | .................... | .................... | .................... |
| H | 401 | hCoV-19/India/TG-CCMB-AB395/2020\|EPI_ISL_539635\|20 | .................... | .................... | .................... | .................... | .................... | .................... |
| H | 402 | hCoV-19/India/TG-CCMB-AA972/2020\|EPI_ISL_539636\|20 | .................... | .................... | .................... | .................... | .................... | .................... |
| H | 403 | hCoV-19/India/TG-CCMB-AB358/2020\|EPI_ISL_539637\|20 | .................... | .................... | .................... | .................... | .................... | .................... |
| H | 404 | hCoV-19/India/TG-CCMB-AA622/2020\|EPI_ISL_539638\|20 | .................... | .................... | .................... | .................... | .................... | .................... |
| H | 405 | hCoV-19/India/TG-CCMB-AB300/2020\|EPI_ISL_539639\|20 | .................... | .................... | .................... | .................... | .................... | .................... |
| H | 406 | hCoV-19/India/TG-CCMB-AB223/2020\|EPI_ISL_539640\|20 | .................... | .................... | .................... | .................... | .................... | .................... |
| H | 407 | hCoV-19/India/TG-CCMB-AA624/2020\|EPI_ISL_539641\|20 | .................... | .................... | .................... | .................... | .................... | .................... |
| H | 408 | hCoV-19/India/TG-CCMB-AC437/2020\|EPI_ISL_539642\|20 | .................... | .................... | .................... | .................... | .................... | .................... |
| H | 409 | hCoV-19/India/TG-CCMB-R305/2020\|EPI_ISL_539643\|202 | .................... | .................... | .................... | .................... | .................... | .................... |
| H | 410 | hCoV-19/India/TG-CCMB-R217/2020\|EPI_ISL_539644\|202 | .................... | .................... | .................... | .................... | .................... | .................... |
| H | 411 | hCoV-19/India/TG-CCMB-AA227/2020\|EPI_ISL_539645\|20 | .................... | .................... | .................... | .................... | .................... | .................... |
| H | 412 | hCoV-19/India/TG-CCMB-AA158/2020\|EPI_ISL_539646\|20 | .................... | .................... | .................... | .................... | .................... | .................... |
| H | 413 | hCoV-19/India/TG-CCMB-AA509/2020\|EPI_ISL_539647\|20 | .................... | .................... | .................... | .................... | .................... | .................... |
| H | 414 | hCoV-19/India/TG-CCMB-R372/2020\|EPI_ISL_539648\|202 | .................... | .................... | .................... | .................... | .................... | .................... |
| H | 415 | hCoV-19/India/TG-CCMB-AA586/2020\|EPI_ISL_539649\|20 | .................... | .................... | .................... | .................... | .................... | .................... |
| H | 416 | hCoV-19/India/TG-CCMB-AA589/2020\|EPI_ISL_539650\|20 | .................... | .................... | .................... | .................... | .................... | .................... |
| H | 417 | hCoV-19/India/TG-CCMB-AA226/2020\|EPI_ISL_539651\|20 | .................... | .................... | .................... | .................... | .................... | .................... |
| H | 418 | hCoV-19/India/TG-CCMB-AA661/2020\|EPI_ISL_539652\|20 | .................... | .................... | .................... | .................... | .................... | .................... |
| H | 419 | hCoV-19/India/TG-CCMB-AA785/2020\|EPI_ISL_539653\|20 | .................... | .................... | .................... | .................... | .................... | .................... |
| H | 420 | hCoV-19/India/TG-CCMB-AA407/2020\|EPI_ISL_539654\|20 | .................... | .................... | .................... | .................... | .................... | .................... |

| | | | | | | | | | |
|---|---|---|---|---|---|---|---|---|---|
| H | 421 | hCoV-19/India/TG-CCMB-AA669/2020\|EPI_ISL_539655\|20 | ···················· | ···················· | ···················· | ···················· | ···················· | ···················· | |
| H | 422 | hCoV-19/India/TG-CCMB-AB314/2020\|EPI_ISL_539656\|20 | ···················· | ···················· | ···················· | ···················· | ···················· | ···················· | |
| H | 423 | hCoV-19/India/TG-CCMB-AB143/2020\|EPI_ISL_539657\|20 | ···················· | ···················· | ···················· | ···················· | ···················· | ···················· | |
| H | 424 | hCoV-19/India/TG-CCMB-AC284/2020\|EPI_ISL_539658\|20 | ···················· | ···················· | ···················· | ···················· | ···················· | ···················· | |
| H | 425 | hCoV-19/India/TG-CCMB-AC434/2020\|EPI_ISL_539659\|20 | ···················· | ···················· | ···················· | ···················· | ···················· | ···················· | |
| H | 426 | hCoV-19/India/TG-CCMB-AC438/2020\|EPI_ISL_539660\|20 | ···················· | ···················· | ···················· | ···················· | ···················· | ···················· | |
| H | 427 | hCoV-19/India/TG-CCMB-AC488/2020\|EPI_ISL_539661\|20 | ···················· | ···················· | ···················· | ···················· | ···················· | ···················· | |
| H | 428 | hCoV-19/India/TG-CCMB-AB336/2020\|EPI_ISL_539662\|20 | ···················· | ···················· | ···················· | ···················· | ···················· | ···················· | |
| H | 429 | hCoV-19/India/TG-CCMB-AA332/2020\|EPI_ISL_539663\|20 | ···················· | ···················· | ···················· | ···················· | ···················· | ···················· | |
| H | 430 | hCoV-19/India/TG-CCMB-AB203/2020\|EPI_ISL_539664\|20 | ···················· | ···················· | ···················· | ···················· | ···················· | ···················· | |
| H | 431 | hCoV-19/India/TG-CCMB-AB578/2020\|EPI_ISL_539665\|20 | ···················· | ···················· | ···················· | ···················· | ···················· | ···················· | |
| H | 432 | hCoV-19/India/TG-CCMB-AB397/2020\|EPI_ISL_539666\|20 | ···················· | ···················· | ···················· | ···················· | ···················· | ···················· | |
| H | 433 | hCoV-19/India/TG-CCMB-AA01/2020\|EPI_ISL_539667\|202 | ···················· | ···················· | ···················· | ···················· | ···················· | ···················· | |
| H | 434 | hCoV-19/India/TG-CCMB-AA33/2020\|EPI_ISL_539668\|202 | ···················· | ···················· | ···················· | ···················· | ···················· | ···················· | |
| H | 435 | hCoV-19/India/TG-CCMB-AA819/2020\|EPI_ISL_539669\|20 | ···················· | ···················· | ···················· | ···················· | ···················· | ···················· | |
| H | 436 | hCoV-19/India/TG-CCMB-AA524/2020\|EPI_ISL_539670\|20 | ···················· | ···················· | ···················· | ···················· | ···················· | ···················· | |
| H | 437 | hCoV-19/India/TG-CCMB-AA863/2020\|EPI_ISL_539671\|20 | ···················· | ···················· | ···················· | ···················· | ···················· | ···················· | |
| H | 438 | hCoV-19/India/TG-CCMB-AA320/2020\|EPI_ISL_539672\|20 | ··················T· | ···················· | ···················· | ···················· | ···················· | ···················· | 1 |
| H | 439 | hCoV-19/India/TG-CCMB-AC377/2020\|EPI_ISL_539673\|20 | ···················· | ···················· | ···················· | ···················· | ···················· | ···················· | |
| H | 440 | hCoV-19/India/TG-CCMB-AA900/2020\|EPI_ISL_539674\|20 | ···················· | ···················· | ···················· | ···················· | ···················· | ···················· | |
| H | 441 | hCoV-19/India/TG-CCMB-AB559/2020\|EPI_ISL_539675\|20 | ···················· | ···················· | ···················· | ···················· | ···················· | ···················· | |
| H | 442 | hCoV-19/India/TG-CCMB-AA04/2020\|EPI_ISL_539676\|202 | ···················· | ···················· | ···················· | ···················· | ···················· | ···················· | |
| H | 443 | hCoV-19/India/TG-CCMB-AA102/2020\|EPI_ISL_539677\|20 | ···················· | ···················· | ···················· | ···················· | ···················· | ···················· | |
| H | 444 | hCoV-19/India/TG-CCMB-AA773/2020\|EPI_ISL_539678\|20 | ···················· | ···················· | ···················· | ···················· | ···················· | ···················· | |
| H | 445 | hCoV-19/India/TG-CCMB-R187/2020\|EPI_ISL_539679\|202 | ···················· | ···················· | ···················· | ···················· | ···················· | ···················· | |
| H | 446 | hCoV-19/India/TG-CCMB-AA19/2020\|EPI_ISL_539680\|202 | ···················· | ···················· | ···················· | ···················· | ···················· | ···················· | |
| H | 447 | hCoV-19/India/TG-CCMB-AB259/2020\|EPI_ISL_539681\|20 | ···················· | ···················· | ···················· | ···················· | ···················· | ···················· | |
| H | 448 | hCoV-19/India/TG-CCMB-AB268/2020\|EPI_ISL_539682\|20 | ···················· | ···················· | ···················· | ···················· | ···················· | ···················· | |
| H | 449 | hCoV-19/India/TG-CCMB-AA105/2020\|EPI_ISL_539683\|20 | ···················· | ···················· | ···················· | ···················· | ···················· | ···················· | |
| H | 450 | hCoV-19/India/TG-CCMB-AA28/2020\|EPI_ISL_539684\|202 | ···················· | ···················· | ···················· | ···················· | ···················· | ···················· | |
| H | 451 | hCoV-19/India/TG-CCMB-AA378/2020\|EPI_ISL_539685\|20 | ···················· | ···················· | ···················· | ···················· | ···················· | ···················· | |
| H | 452 | hCoV-19/India/TG-CCMB-AA608/2020\|EPI_ISL_539686\|20 | ···················· | ···················· | ···················· | ···················· | ···················· | ···················· | |
| H | 453 | hCoV-19/India/TG-CCMB-AA370/2020\|EPI_ISL_539687\|20 | ···················· | ···················· | ···················· | ···················· | ···················· | ···················· | |
| H | 454 | hCoV-19/India/TG-CCMB-AA371/2020\|EPI_ISL_539688\|20 | ···················· | ···················· | ···················· | ···················· | ···················· | ···················· | |
| H | 455 | hCoV-19/India/TG-CCMB-AA375/2020\|EPI_ISL_539689\|20 | ···················· | ···················· | ···················· | ···················· | ···················· | ···················· | |
| H | 456 | hCoV-19/England/LIVE-AABE9/2020\|EPI_ISL_540603\|202 | ···················· | ···················· | ···················· | ···················· | ···················· | ···················· | |
| H | 459 | hCoV-19/England/LIVE-AACC8/2020\|EPI_ISL_540610\|202 | ···················· | ···················· | ···················· | ···················· | ···················· | ···················· | |
| H | 461 | hCoV-19/England/LIVE-AAD01/2020\|EPI_ISL_540614\|202 | ···················· | ···················· | ···················· | ···················· | ···················· | ···················· | |
| H | 463 | hCoV-19/England/LIVE-AAE0E/2020\|EPI_ISL_540618\|202 | ···················· | ···················· | ···················· | ···················· | ···················· | ···················· | |
| H | 464 | hCoV-19/England/LIVE-AAE1D/2020\|EPI_ISL_540619\|202 | ···················· | ···················· | ···················· | ···················· | ···················· | ···················· | |
| H | 466 | hCoV-19/England/LIVE-AAE95/2020\|EPI_ISL_540622\|202 | ···················· | ···················· | ···················· | ···················· | ···················· | ···················· | |
| H | 467 | hCoV-19/England/LIVE-AAEA4/2020\|EPI_ISL_540623\|202 | ···················· | ···················· | ···················· | ···················· | ···················· | ···················· | |
| H | 468 | hCoV-19/England/LIVE-AAF47/2020\|EPI_ISL_540624\|202 | ···················· | ···················· | ···················· | ···················· | ···················· | ···················· | |
| H | 470 | hCoV-19/England/LIVE-ABCB8/2020\|EPI_ISL_540629\|202 | ···················· | ···················· | ···················· | ···················· | ···················· | ···················· | |
| H | 471 | hCoV-19/England/MILK-991F95/2020\|EPI_ISL_540640\|20 | ···················· | ···················· | ···················· | ···················· | ···················· | ···················· | |
| H | 472 | hCoV-19/England/MILK-991FB3/2020\|EPI_ISL_540641\|20 | ···················· | ···················· | ···················· | ···················· | ···················· | ···················· | |
| H | 473 | hCoV-19/England/MILK-991FFF/2020\|EPI_ISL_540642\|20 | ···················· | ···················· | ···················· | ···················· | ···················· | ···················· | |
| H | 474 | hCoV-19/England/MILK-99200A/2020\|EPI_ISL_540643\|20 | ···················· | ···················· | ···················· | ···················· | ···················· | ···················· | |
| H | 475 | hCoV-19/England/MILK-992046/2020\|EPI_ISL_540645\|20 | ···················· | ···················· | ···················· | ···················· | ···················· | ···················· | |
| H | 476 | hCoV-19/England/MILK-992064/2020\|EPI_ISL_540646\|20 | ···················· | ·A·················· | ···················· | ···················· | ···················· | ···················· | |
| H | 477 | hCoV-19/England/MILK-9920BF/2020\|EPI_ISL_540647\|20 | ···················· | ···················· | ···················· | ···················· | ···················· | ···················· | |
| H | 478 | hCoV-19/England/MILK-9920EC/2020\|EPI_ISL_540648\|20 | ···················· | ·A·················· | ···················· | ···················· | ···················· | ··A················· | |
| H | 479 | hCoV-19/England/MILK-992125/2020\|EPI_ISL_540649\|20 | ···················· | ·A·················· | ···················· | ···················· | ···················· | ··A················· | |
| H | 480 | hCoV-19/England/MILK-992152/2020\|EPI_ISL_540650\|20 | ···················· | ···················· | ···················· | ···················· | ···················· | ···················· | |
| H | 481 | hCoV-19/England/MILK-992161/2020\|EPI_ISL_540651\|20 | ···················· | ···················· | ···················· | ···················· | ···················· | ···················· | |
| H | 482 | hCoV-19/England/MILK-992170/2020\|EPI_ISL_540652\|20 | ···················· | ···················· | ···················· | ···················· | ···················· | ···················· | |
| H | 483 | hCoV-19/England/MILK-9921CB/2020\|EPI_ISL_540656\|20 | ···················· | ·A·················· | ···················· | ···················· | ···················· | ··A················· | |
| H | 484 | hCoV-19/England/MILK-9921E9/2020\|EPI_ISL_540658\|20 | ···················· | ···················· | ···················· | ···················· | ···················· | ···················· | |
| H | 485 | hCoV-19/England/MILK-992213/2020\|EPI_ISL_540659\|20 | ···················· | ···················· | ···················· | ···················· | ···················· | ···················· | |
| H | 486 | hCoV-19/England/MILK-992222/2020\|EPI_ISL_540660\|20 | ···················· | ···················· | ···················· | ···················· | ···················· | ···················· | |
| H | 487 | hCoV-19/England/MILK-992231/2020\|EPI_ISL_540661\|20 | ···················· | ·A·················· | ···················· | ···················· | ···················· | ··A················· | |
| H | 488 | hCoV-19/England/MILK-99225F/2020\|EPI_ISL_540663\|20 | ···················· | ···················· | ···················· | ···················· | ···················· | ···················· | |
| H | 489 | hCoV-19/England/MILK-99227D/2020\|EPI_ISL_540664\|20 | ···················· | ···················· | ···················· | ···················· | ···················· | ···················· | |
| H | 490 | hCoV-19/England/MILK-9922B9/2020\|EPI_ISL_540667\|20 | ···················· | ···················· | ···················· | ···················· | ···················· | ···················· | |
| H | 491 | hCoV-19/England/MILK-9922C8/2020\|EPI_ISL_540668\|20 | ···················· | ···················· | ···················· | ···················· | ···················· | ···················· | |
| H | 492 | hCoV-19/England/MILK-99232F/2020\|EPI_ISL_540672\|20 | ···················· | ···················· | ···················· | ···················· | ···················· | ···················· | |
| H | 493 | hCoV-19/England/MILK-99234D/2020\|EPI_ISL_540673\|20 | ···················· | ···················· | ···················· | ···················· | ···················· | ···················· | |
| H | 494 | hCoV-19/England/MILK-9923B6/2020\|EPI_ISL_540676\|20 | ···················· | ···················· | ···················· | ···················· | ···················· | ···················· | |
| H | 495 | hCoV-19/England/MILK-9923E3/2020\|EPI_ISL_540678\|20 | ···················· | ···················· | ···················· | ···················· | ···················· | ···················· | |
| H | 496 | hCoV-19/England/MILK-9923F2/2020\|EPI_ISL_540679\|20 | ···················· | ···················· | ···················· | ···················· | ···················· | ···················· | |
| H | 498 | hCoV-19/Spain/MD-IBV-99007164/2020\|EPI_ISL_541934\| | ···················· | ···················· | ···················· | ···················· | ···················· | ···················· | |
| H | 501 | hCoV-19/Spain/MD-IBV-99007169/2020\|EPI_ISL_541939\| | ···················· | ···················· | ···················· | ···················· | ···················· | ···················· | |
| H | 512 | hCoV-19/Norway/Bergen-01/2020\|EPI_ISL_541970\|2020- | ···················· | ···················· | ···················· | ···················· | ···················· | ···················· | |
| H | 513 | hCoV-19/Northern Ireland/QEUH-960DA4/2020\|EPI_ISL_ | ···················· | ···················· | ···················· | ···················· | ···················· | ···················· | |
| H | 514 | hCoV-19/England/QEUH-960955/2020\|EPI_ISL_532029\|20 | ···················· | ···················· | ···················· | ···················· | ···················· | ···················· | |
| H | 515 | hCoV-19/Wales/QEUH-961040/2020\|EPI_ISL_532030\|2020 | ···················· | ···················· | ···················· | ···················· | ···················· | ···················· | |
| H | 516 | hCoV-19/Northern Ireland/QEUH-960876/2020\|EPI_ISL_ | ···················· | ···················· | ···················· | ···················· | ···················· | ···················· | |
| H | 517 | hCoV-19/England/QEUH-960C7A/2020\|EPI_ISL_532033\|20 | ···················· | ···················· | ···················· | ···················· | ···················· | ···················· | |
| H | 518 | hCoV-19/England/QEUH-9605F7/2020\|EPI_ISL_532034\|20 | ···················· | ···················· | ···················· | ···················· | ···················· | ···················· | |
| H | 519 | hCoV-19/England/QEUH-96108C/2020\|EPI_ISL_532035\|20 | ···················· | ···················· | ···················· | ···················· | ···················· | ···················· | |

```
H 520 hCoV-19/England/QEUH-96113E/2020|EPI_ISL_532036|20 ···················· ···················· ····················· ···················· ···················· ····················
H 521 hCoV-19/England/QEUH-960AF8/2020|EPI_ISL_532037|20 ···················· ···················· ····················· ···················· ···················· ····················
H 522 hCoV-19/Scotland/QEUH-96114D/2020|EPI_ISL_532038|2 ···················· ···················· ····················· ···················· ···················· ····················
H 523 hCoV-19/Northern Ireland/QEUH-960E47/2020|EPI_ISL_ ···················· ···················· ····················· ···················· ···················· ····················
H 524 hCoV-19/England/QEUH-960603/2020|EPI_ISL_532040|20 ···················· ···················· ····················· ···················· ···················· ····················
H 525 hCoV-19/England/QEUH-96015D/2020|EPI_ISL_532041|20 ···················· ···················· ····················· ···················· ···················· ····················
H 526 hCoV-19/England/QEUH-96106E/2020|EPI_ISL_532042|20 ···················· ···················· ····················· ···················· ···················· ····················
H 527 hCoV-19/England/QEUH-9608FE/2020|EPI_ISL_532043|20 ···················· ···················· ····················· ···················· ···················· ····················
H 528 hCoV-19/England/QEUH-960894/2020|EPI_ISL_532044|20 ···················· ···················· ····················· ···················· ···················· ····················
H 529 hCoV-19/England/QEUH-96065E/2020|EPI_ISL_532045|20 ···················· ···················· ····················· ···················· ···················· ····················
H 530 hCoV-19/England/QEUH-96064F/2020|EPI_ISL_532046|20 ···················· ···················· ····················· ···················· ···················· ····················
H 531 hCoV-19/England/QEUH-960524/2020|EPI_ISL_532047|20 ···················· ···················· ····················· ···················· ···················· ····················
H 532 hCoV-19/Scotland/QEUH-960542/2020|EPI_ISL_532049|2 ···················· ···················· ····················· ···················· ···················· ····················
H 533 hCoV-19/England/QEUH-960B7D/2020|EPI_ISL_532050|20 ···················· ···················· ····················· ···················· ···················· ····················
H 534 hCoV-19/England/QEUH-960A43/2020|EPI_ISL_532052|20 ···················· ···················· ····················· ···················· ···················· ····················
H 535 hCoV-19/Scotland/QEUH-960515/2020|EPI_ISL_532053|2 ···················· ···················· ····················· ···················· ···················· ····················
H 536 hCoV-19/England/QEUH-9604EB/2020|EPI_ISL_532054|20 ···················· ···················· ····················· ···················· ···················· ····················
H 537 hCoV-19/Scotland/QEUH-9607B5/2020|EPI_ISL_532055|2 ···················· ···················· ····················· ···················· ···················· ····················
H 538 hCoV-19/England/QEUH-9606C7/2020|EPI_ISL_532056|20 ···················· ···················· ····················· ···················· ···················· ····················
H 539 hCoV-19/Scotland/QEUH-961347/2020|EPI_ISL_532057|2 ···················· ···················· ····················· ···················· ···················· ····················
H 540 hCoV-19/England/QEUH-9612C2/2020|EPI_ISL_532058|20 ···················· ···················· ····················· ···················· ···················· ····················
H 541 hCoV-19/England/QEUH-96074C/2020|EPI_ISL_532059|20 ···················· ···················· ····················· ···················· ···················· ····················
H 542 hCoV-19/Northern Ireland/QEUH-9607D3/2020|EPI_ISL_ ···················· ···················· ····················· ···················· ···················· ····················
H 543 hCoV-19/Scotland/QEUH-9606F4/2020|EPI_ISL_532062|2 ···················· ···················· ····················· ···················· ···················· ····················
H 544 hCoV-19/England/QEUH-960788/2020|EPI_ISL_532063|20 ···················· ···················· ····················· ···················· ···················· ····················
H 545 hCoV-19/England/QEUH-960481/2020|EPI_ISL_532064|20 ···················· ···················· ····················· ···················· ···················· ····················
H 546 hCoV-19/England/QEUH-9609EC/2020|EPI_ISL_532065|20 ···················· ···················· ····················· ···················· ···················· ····················
H 547 hCoV-19/England/QEUH-960B5F/2020|EPI_ISL_532066|20 ···················· ···················· ····················· ···················· ···················· ····················
H 548 hCoV-19/England/QEUH-960BB9/2020|EPI_ISL_532067|20 ···················· ···················· ····················· ···················· ···················· ····················
H 549 hCoV-19/England/QEUH-9602B4/2020|EPI_ISL_532068|20 ···················· ···················· ····················· ···················· ···················· ····················
H 550 hCoV-19/England/QEUH-960F08/2020|EPI_ISL_532069|20 ···················· ·A·················· ····················· ···················· ···················· ····················
H 551 hCoV-19/Scotland/QEUH-96115C/2020|EPI_ISL_532070|2 ···················· ···················· ····················· ···················· ···················· ····················
H 552 hCoV-19/England/QEUH-9604FA/2020|EPI_ISL_532072|20 ···················· ···················· ····················· ···················· ···················· ····················
H 553 hCoV-19/England/QEUH-960B8C/2020|EPI_ISL_532073|20 ···················· ·A·················· ····················· ···················· ···················· ····················
H 554 hCoV-19/England/QEUH-960937/2020|EPI_ISL_532074|20 ···················· ···················· ····················· ···················· ···················· ····················
H 555 hCoV-19/England/QEUH-9608EF/2020|EPI_ISL_532075|20 ···················· ···················· ····················· ···················· ···················· ····················
H 556 hCoV-19/Scotland/QEUH-96122C/2020|EPI_ISL_532076|2 ···················· ···················· ····················· ···················· ···················· ····················
H 557 hCoV-19/Scotland/QEUH-96130B/2020|EPI_ISL_532077|2 ···················· ···················· ····················· ···················· ···················· ····················
H 558 hCoV-19/Scotland/QEUH-960CF2/2020|EPI_ISL_532078|2 ···················· ···················· ····················· ···················· ···················· ····················
H 559 hCoV-19/England/QEUH-96024B/2020|EPI_ISL_532079|20 ···················· ···················· ····················· ···················· ····T··············· ····················
H 560 hCoV-19/Scotland/QEUH-961198/2020|EPI_ISL_532080|2 ···················· ···················· ····················· ···················· ···················· ····················
H 561 hCoV-19/Scotland/QEUH-9611B6/2020|EPI_ISL_532081|2 ···················· ···················· ····················· ···················· ···················· ····················
H 562 hCoV-19/England/QEUH-961101/2020|EPI_ISL_532082|20 ···················· ···················· ····················· ···················· ···················· ····················
H 563 hCoV-19/England/QEUH-96068B/2020|EPI_ISL_532083|20 ···················· ···················· ····················· ···················· ···················· ····················
H 564 hCoV-19/England/QEUH-960700/2020|EPI_ISL_532084|20 ···················· ···················· ····················· ···················· ···················· ····················
H 565 hCoV-19/England/QEUH-960D4A/2020|EPI_ISL_532086|20 ···················· ·A·················· ····················· ···················· ···················· ···A················
H 566 hCoV-19/Northern Ireland/QEUH-960F26/2020|EPI_ISL_ ···················· ···················· ····················· ···················· ···················· ····················
H 567 hCoV-19/England/QEUH-960384/2020|EPI_ISL_532088|20 ···················· ···················· ····················· ···················· ···················· ····················
H 568 hCoV-19/England/QEUH-9612D1/2020|EPI_ISL_532089|20 ···················· ···················· ····················· ···················· ···················· ····················
H 569 hCoV-19/England/QEUH-96124A/2020|EPI_ISL_532090|20 ···················· ···················· ····················· ···················· ···················· ····················
H 570 hCoV-19/England/QEUH-960393/2020|EPI_ISL_532091|20 ···················· ···················· ····················· ···················· ···················· ····················
H 571 hCoV-19/England/QEUH-96025A/2020|EPI_ISL_532092|20 ···················· ···················· ····················· ···················· ···················· ····················
H 572 hCoV-19/Scotland/QEUH-960C01/2020|EPI_ISL_532093|2 ···················· ···················· ····················· ···················· ···················· ····················
H 573 hCoV-19/Scotland/QEUH-9611C5/2020|EPI_ISL_532094|2 ···················· ···················· ····················· ···················· ···················· ····················
H 574 hCoV-19/England/QEUH-9602C3/2020|EPI_ISL_532095|20 ···················· ···················· ····················· ···················· ···················· ····················
H 575 hCoV-19/England/QEUH-9610E6/2020|EPI_ISL_532096|20 ···················· ···················· ····················· ···················· ···················· ····················
H 576 hCoV-19/England/QEUH-96032A/2020|EPI_ISL_532097|20 ···················· ···················· ····················· ···················· ····T··············· ····················
H 577 hCoV-19/Scotland/QEUH-9610D7/2020|EPI_ISL_532098|2 ···················· ···················· ····················· ···················· ···················· ····················
H 578 hCoV-19/England/QEUH-9606B8/2020|EPI_ISL_532099|20 ···················· ···················· ····················· ···················· ···················· ····················
H 579 hCoV-19/England/QEUH-5E62AB/2020|EPI_ISL_533300|20 ···················· ···················· ····················· ···················· ···················· ····················
H 580 hCoV-19/England/QEUH-5EC4BE/2020|EPI_ISL_533301|20 ···················· ···················· ····················· ···················· ···················· ····················
H 581 hCoV-19/England/QEUH-5E004D/2020|EPI_ISL_533302|20 ···················· ···················· ····················· ···················· ···················· ····················
H 582 hCoV-19/England/QEUH-5EAB06/2020|EPI_ISL_533303|20 ···················· ···················· ····················· ···················· ···················· ····················
H 583 hCoV-19/England/QEUH-5E93F0/2020|EPI_ISL_533304|20 ···················· ···················· ····················· ···················· ···················· ····················
H 584 hCoV-19/Northern Ireland/QEUH-5E542E/2020|EPI_ISL_ ···················· ······A············· ····················· ···················· ···················· ····················
H 585 hCoV-19/England/QEUH-5DB4FE/2020|EPI_ISL_533306|20 ···················· ···················· ····················· ···················· ···················· ····················
H 586 hCoV-19/England/QEUH-5E65B1/2020|EPI_ISL_533308|20 ···················· ···················· ····················· ···················· ···················· ····················
H 587 hCoV-19/England/QEUH-5DD5BD/2020|EPI_ISL_533309|20 ···················· ···················· ····················· ···················· ···················· ····················
H 588 hCoV-19/England/QEUH-5DD18C/2020|EPI_ISL_533310|20 ···················· ···················· ····················· ···················· ···················· ····················
H 589 hCoV-19/Scotland/QEUH-5DCCB9/2020|EPI_ISL_533311|2 ···················· ···················· ····················· ···················· ···················· ····················
H 590 hCoV-19/England/QEUH-5E778D/2020|EPI_ISL_533313|20 ···················· ···················· ····················· ···················· ···················· ····················
H 591 hCoV-19/England/QEUH-5E751A/2020|EPI_ISL_533314|20 ···················· ···················· ····················· ···················· ···················· ····················
H 592 hCoV-19/England/QEUH-5E8D20/2020|EPI_ISL_533315|20 ···················· ···················· ····················· ···················· ···················· ····················
H 593 hCoV-19/Northern Ireland/QEUH-5E5330/2020|EPI_ISL_ ···················· ···················· ····················· ···················· ···················· ····················
H 594 hCoV-19/England/QEUH-5E7B72/2020|EPI_ISL_533318|20 ···················· ···················· ····················· ···················· ···················· ····················
H 595 hCoV-19/England/QEUH-5E4559/2020|EPI_ISL_533319|20 ···················· ···A················ ····················· ···················· ···················· ····················
H 596 hCoV-19/England/QEUH-5EB58F/2020|EPI_ISL_533320|20 ···················· ···················· ····················· ···················· ···················· ····················
H 597 hCoV-19/England/QEUH-5DE4A0/2020|EPI_ISL_533321|20 ···················· ···················· ····················· ···················· ···················· ····················
H 598 hCoV-19/England/QEUH-5E8537/2020|EPI_ISL_533323|20 ···················· ···················A ····················· ···················· ···················· ····················  1
H 599 hCoV-19/England/QEUH-542F97/2020|EPI_ISL_533360|20 ···················· ···················· ····················· ···················· ···················· ····················
```

| | | | | | | | | | |
|---|---|---|---|---|---|---|---|---|---|
| H | 600 | hCoV-19/England/QEUH-5460BE/2020\|EPI_ISL_533361\|20 | .................... | .................... | .................... | .................... | .................... | .................... | |
| H | 601 | hCoV-19/England/QEUH-543A95/2020\|EPI_ISL_533362\|20 | .................... | .................... | .................... | .................... | .................... | .................... | |
| H | 602 | hCoV-19/Wales/QEUH-543075/2020\|EPI_ISL_533363\|2020 | .................... | .................... | .................... | .................... | .................... | .................... | |
| H | 603 | hCoV-19/England/QEUH-544733/2020\|EPI_ISL_533364\|20 | .................... | .A.................. | .................... | .................... | .................... | .................... | |
| H | 604 | hCoV-19/England/QEUH-545CD9/2020\|EPI_ISL_533366\|20 | .................... | .................... | .................... | .................... | .................... | .................... | |
| H | 605 | hCoV-19/England/QEUH-54590F/2020\|EPI_ISL_533368\|20 | .................... | .................... | .................... | .................... | .................... | .................... | |
| H | 606 | hCoV-19/England/QEUH-54D5C7/2020\|EPI_ISL_533369\|20 | .................... | .................... | .................... | .................... | .................... | .................... | |
| H | 607 | hCoV-19/England/QEUH-5475AF/2020\|EPI_ISL_533370\|20 | .................... | .................... | .................... | .................... | .................... | .................... | |
| H | 608 | hCoV-19/England/QEUH-54933A/2020\|EPI_ISL_533371\|20 | .................... | ...................A | .................... | .................... | .................... | .................... | 1 |
| H | 609 | hCoV-19/England/QEUH-54B8A2/2020\|EPI_ISL_533372\|20 | .................... | .................... | .................... | .................... | .................... | .................... | |
| H | 610 | hCoV-19/England/QEUH-54D5D6/2020\|EPI_ISL_533374\|20 | .................... | .................... | .................... | .................... | .................... | .................... | |
| H | 611 | hCoV-19/England/QEUH-54D54F/2020\|EPI_ISL_533375\|20 | .................... | .................... | .................... | .................... | .................... | .................... | |
| H | 612 | hCoV-19/Scotland/QEUH-5484A1/2020\|EPI_ISL_533376\|2 | .................... | ...................A | .................... | .................... | .................... | .................... | 1 |
| H | 613 | hCoV-19/England/QEUH-5477A9/2020\|EPI_ISL_533377\|20 | .................... | .................... | .................... | .................... | .................... | .................... | |
| H | 614 | hCoV-19/England/QEUH-54A339/2020\|EPI_ISL_533378\|20 | .................... | .................... | .................... | .................... | .................... | .................... | |
| H | 615 | hCoV-19/England/QEUH-54D433/2020\|EPI_ISL_533379\|20 | .................... | .................... | .................... | .................... | .................... | .................... | |
| H | 616 | hCoV-19/England/QEUH-546713/2020\|EPI_ISL_533380\|20 | .................... | .................... | .................... | .................... | .................... | .................... | |
| H | 617 | hCoV-19/England/QEUH-54B73C/2020\|EPI_ISL_533381\|20 | .................... | .................... | .................... | .................... | .................... | .................... | |
| H | 618 | hCoV-19/England/QEUH-54D7D0/2020\|EPI_ISL_533382\|20 | .................... | .................... | .................... | .................... | .................... | .................... | |
| H | 619 | hCoV-19/England/QEUH-54CC69/2020\|EPI_ISL_533383\|20 | .................... | .................... | .................... | .................... | .................... | .................... | |
| H | 620 | hCoV-19/England/QEUH-548298/2020\|EPI_ISL_533384\|20 | .................... | .................... | .................... | .................... | .................... | .................... | |
| H | 621 | hCoV-19/England/QEUH-54AA34/2020\|EPI_ISL_533385\|20 | .................... | .................... | .................... | .................... | .................... | .................... | |
| H | 622 | hCoV-19/England/QEUH-54B81B/2020\|EPI_ISL_533386\|20 | .................... | .................... | .................... | .................... | .................... | .................... | |
| H | 623 | hCoV-19/England/QEUH-54CBF3/2020\|EPI_ISL_533388\|20 | .................... | .................... | .................... | .................... | .................... | .................... | |
| H | 624 | hCoV-19/England/QEUH-54CC78/2020\|EPI_ISL_533389\|20 | .................... | .................... | .................... | .................... | .................... | .................... | |
| H | 625 | hCoV-19/England/QEUH-5476CA/2020\|EPI_ISL_533390\|20 | .................... | .................... | .................... | .................... | .................... | .................... | |
| H | 626 | hCoV-19/England/QEUH-549710/2020\|EPI_ISL_533391\|20 | .................... | .................... | .................... | .................... | .................... | .................... | |
| H | 627 | hCoV-19/England/QEUH-54DE08/2020\|EPI_ISL_533392\|20 | .................... | .................... | .................... | .................... | .................... | .................... | |
| H | 628 | hCoV-19/England/QEUH-5466E9/2020\|EPI_ISL_533393\|20 | .................... | .A.................. | .................... | .................... | .................... | .................... | |
| H | 629 | hCoV-19/Scotland/QEUH-54BF25/2020\|EPI_ISL_533394\|2 | .................... | .................... | .................... | .................... | .................... | .................... | |
| H | 630 | hCoV-19/England/QEUH-549604/2020\|EPI_ISL_533395\|20 | .................... | .................... | .................... | .................... | .................... | .................... | |
| H | 631 | hCoV-19/England/QEUH-5482C5/2020\|EPI_ISL_533397\|20 | .................... | .................... | .................... | .................... | .................... | .................... | |
| H | 632 | hCoV-19/England/QEUH-54DDB0/2020\|EPI_ISL_533398\|20 | .................... | .................... | .................... | .................... | .................... | .................... | |
| H | 633 | hCoV-19/Scotland/QEUH-96CF0C/2020\|EPI_ISL_534602\|2 | .................... | .................... | .................... | .................... | .................... | .................... | |
| H | 634 | hCoV-19/Scotland/QEUH-96D7D6/2020\|EPI_ISL_534603\|2 | .................... | .................... | .................... | .................... | .................... | .................... | |
| H | 635 | hCoV-19/Scotland/QEUH-96D75E/2020\|EPI_ISL_534605\|2 | .................... | .................... | .................... | .................... | .................... | .................... | |
| H | 636 | hCoV-19/Scotland/QEUH-96D730/2020\|EPI_ISL_534606\|2 | .................... | .................... | .................... | .................... | .................... | .................... | |
| H | 637 | hCoV-19/Scotland/QEUH-96D509/2020\|EPI_ISL_534607\|2 | .................... | .................... | .................... | .................... | .................... | .................... | |
| H | 638 | hCoV-19/Scotland/QEUH-96C7AA/2020\|EPI_ISL_534608\|2 | .................... | .................... | .................... | .................... | .................... | .................... | |
| H | 639 | hCoV-19/Scotland/QEUH-96D4EE/2020\|EPI_ISL_534609\|2 | .................... | .................... | .................... | .................... | .................... | .................... | |
| H | 640 | hCoV-19/Scotland/QEUH-96D76D/2020\|EPI_ISL_534610\|2 | .................... | .................... | .................... | .................... | .................... | .................... | |
| H | 641 | hCoV-19/Scotland/QEUH-96D527/2020\|EPI_ISL_534611\|2 | .................... | .................... | .................... | .................... | .................... | .................... | |
| H | 642 | hCoV-19/Scotland/QEUH-96D5CD/2020\|EPI_ISL_534613\|2 | .................... | .................... | .................... | .................... | .................... | .................... | |
| H | 643 | hCoV-19/Scotland/QEUH-96D0F9/2020\|EPI_ISL_534614\|2 | .................... | .................... | .................... | .................... | .................... | .................... | |
| H | 644 | hCoV-19/Scotland/QEUH-96D35A/2020\|EPI_ISL_534615\|2 | .................... | .................... | .................... | .................... | .................... | .................... | |
| H | 645 | hCoV-19/Scotland/QEUH-96CF57/2020\|EPI_ISL_534616\|2 | .................... | .................... | .................... | .................... | .................... | .................... | |
| H | 646 | hCoV-19/Scotland/QEUH-96C77D/2020\|EPI_ISL_534617\|2 | .................... | .A.................. | .................... | .................... | .................... | .................... | |
| H | 647 | hCoV-19/Scotland/QEUH-96D7B8/2020\|EPI_ISL_534619\|2 | .................... | .................... | .................... | .................... | .................... | .................... | |
| H | 648 | hCoV-19/Scotland/QEUH-96D518/2020\|EPI_ISL_534620\|2 | .................... | .................... | .................... | .................... | .................... | .................... | |
| H | 649 | hCoV-19/Scotland/QEUH-96C889/2020\|EPI_ISL_534621\|2 | .................... | .................... | .................... | .................... | .................... | .................... | |
| H | 650 | hCoV-19/Scotland/QEUH-96CF2A/2020\|EPI_ISL_534623\|2 | .................... | .................... | .................... | .................... | .................... | .................... | |
| H | 651 | hCoV-19/Scotland/QEUH-96CDC6/2020\|EPI_ISL_534624\|2 | .................... | .................... | .................... | .................... | .................... | .................... | |
| H | 652 | hCoV-19/Scotland/QEUH-96D4FD/2020\|EPI_ISL_534625\|2 | .................... | .................... | .................... | .................... | .................... | .................... | |
| H | 653 | hCoV-19/Scotland/QEUH-96D6BB/2020\|EPI_ISL_534627\|2 | .................... | .................... | .................... | .................... | .................... | .................... | |
| H | 654 | hCoV-19/Scotland/QEUH-96D67F/2020\|EPI_ISL_534628\|2 | .................... | .................... | .................... | .................... | .................... | .................... | |
| H | 655 | hCoV-19/Scotland/QEUH-96CD20/2020\|EPI_ISL_534629\|2 | .................... | .................... | .................... | .................... | .................... | .................... | |
| H | 656 | hCoV-19/Scotland/QEUH-96D572/2020\|EPI_ISL_534630\|2 | .................... | .................... | .................... | .................... | .................... | .................... | |
| H | 657 | hCoV-19/Scotland/QEUH-96C898/2020\|EPI_ISL_534631\|2 | .................... | .................... | .................... | .................... | .................... | .................... | |
| H | 658 | hCoV-19/Scotland/QEUH-96D23F/2020\|EPI_ISL_534632\|2 | .................... | .................... | .................... | .................... | .................... | .................... | |
| H | 659 | hCoV-19/Scotland/QEUH-96CC05/2020\|EPI_ISL_534633\|2 | .................... | .................... | .................... | .................... | .................... | .................... | |
| H | 660 | hCoV-19/Scotland/QEUH-96C7D7/2020\|EPI_ISL_534636\|2 | .................... | .................... | .................... | .................... | .................... | .................... | |
| H | 661 | hCoV-19/Scotland/QEUH-96D32D/2020\|EPI_ISL_534637\|2 | .................... | .................... | .................... | .................... | .................... | .................... | |
| H | 662 | hCoV-19/Scotland/QEUH-96CF66/2020\|EPI_ISL_534638\|2 | .................... | .................... | .................... | .................... | .................... | .................... | |
| H | 663 | hCoV-19/Scotland/QEUH-96C82F/2020\|EPI_ISL_534639\|2 | .................... | .................... | .................... | .................... | .................... | .................... | |
| H | 664 | hCoV-19/Scotland/QEUH-96D0BD/2020\|EPI_ISL_534640\|2 | .................... | .................... | .................... | .................... | .................... | .................... | |
| H | 665 | hCoV-19/Scotland/QEUH-96D6F7/2020\|EPI_ISL_534641\|2 | .................... | .................... | .................... | .................... | .................... | .................... | |
| H | 666 | hCoV-19/Scotland/QEUH-96CF39/2020\|EPI_ISL_534642\|2 | .................... | .................... | .................... | .................... | .................... | .................... | |
| H | 667 | hCoV-19/Scotland/QEUH-964B3D/2020\|EPI_ISL_534643\|2 | .................... | .................... | .................... | .................... | .................... | .................... | |
| H | 668 | hCoV-19/Scotland/QEUH-96B240/2020\|EPI_ISL_534645\|2 | .................... | .................... | .................... | .................... | .................... | .................... | |
| H | 669 | hCoV-19/Scotland/QEUH-96B36B/2020\|EPI_ISL_534646\|2 | .................... | .................... | .................... | .................... | .................... | .................... | |
| H | 670 | hCoV-19/Scotland/QEUH-96B7AB/2020\|EPI_ISL_534647\|2 | .................... | .................... | .................... | .................... | .................... | .................... | |
| H | 671 | hCoV-19/Scotland/QEUH-96B741/2020\|EPI_ISL_534648\|2 | .................... | .................... | .................... | .................... | .................... | .................... | |
| H | 672 | hCoV-19/Scotland/QEUH-96A997/2020\|EPI_ISL_534650\|2 | ........T........... | .................... | .................... | .................... | .................... | .................... | |
| H | 673 | hCoV-19/Scotland/QEUH-96B398/2020\|EPI_ISL_534651\|2 | .................... | .A.................. | .................... | .................... | .................... | .................... | |
| H | 674 | hCoV-19/Scotland/QEUH-96B3A7/2020\|EPI_ISL_534652\|2 | .................... | .................... | .................... | .................... | .................... | .................... | |
| H | 675 | hCoV-19/Scotland/QEUH-96B538/2020\|EPI_ISL_534690\|2 | .................... | .................... | .................... | .................... | .................... | .................... | |
| H | 676 | hCoV-19/USA/PR-B35B/2020\|EPI_ISL_534693\|2020-08-05 | .................... | .................... | .................... | .................... | .................... | .................... | |
| H | 677 | hCoV-19/USA/PR-B3SB/2020\|EPI_ISL_534694\|2020-08-06 | .................... | .................... | .................... | .................... | .................... | .................... | |
| H | 678 | hCoV-19/USA/PR-B412/2020\|EPI_ISL_534695\|2020-08-06 | .................... | .................... | .................... | .................... | .................... | .................... | |
| H | 679 | hCoV-19/USA/PR-B5XD/2020\|EPI_ISL_534696\|2020-08-11 | .................... | .................... | .................... | .................... | .................... | .................... | |

| | | | | | | | |
|---|---|---|---|---|---|---|---|
| H | 680 hCoV-19/USA/PR-B5XN/2020\|EPI_ISL_534697\|2020-08-11 | ···················· | ···················· | ····················· | ···················· | ···················· | ···················· |
| H | 681 hCoV-19/USA/PR-B6DU/2020\|EPI_ISL_534698\|2020-08-11 | ···················· | ···················· | ····················· | ···················· | ···················· | ···················· |
| H | 682 hCoV-19/USA/MD-MDH-0135/2020\|EPI_ISL_534699\|2020-0 | ···················· | ···················· | ····················· | ···················· | ···················· | ···················· |
| H | 683 hCoV-19/Canada/QC-LSPQ-L00230798/2020\|EPI_ISL_5359 | ···················· | ···················· | ····················· | ···················· | ···················· | ···················· |
| H | 684 hCoV-19/Canada/QC-LSPQ-L00230813/2020\|EPI_ISL_5359 | ···················· | ···················· | ····················· | ···················· | ···················· | ···················· |
| H | 685 hCoV-19/Canada/QC-LSPQ-L00230821/2020\|EPI_ISL_5359 | ···················· | ···················· | ····················· | ···················· | ···················· | ···················· |
| H | 686 hCoV-19/Canada/QC-LSPQ-L00230881/2020\|EPI_ISL_5359 | ···················· | ···················· | ····················· | ···················· | ···················· | ···················· |
| H | 687 hCoV-19/Canada/QC-LSPQ-L00230882/2020\|EPI_ISL_5359 | ···················· | ···················· | ····················· | ···················· | ···················· | ···················· |
| H | 688 hCoV-19/Canada/QC-LSPQ-L00230895/2020\|EPI_ISL_5359 | ···················· | ···················· | ····················· | ···················· | ···················· | ···················· |
| H | 689 hCoV-19/Canada/QC-LSPQ-L00230948/2020\|EPI_ISL_5359 | ···················· | ···················· | ····················· | ···················· | ···················· | ···················· |
| H | 690 hCoV-19/Canada/QC-LSPQ-L00230998/2020\|EPI_ISL_5359 | ···················· | ···················· | ····················· | ···················· | ···················· | ···················· |
| H | 691 hCoV-19/Canada/QC-LSPQ-L00231066/2020\|EPI_ISL_5359 | ···················· | ···················· | ····················· | ···················· | ···················· | ···················· |
| H | 692 hCoV-19/Canada/QC-LSPQ-L00231068/2020\|EPI_ISL_5359 | ···················· | ···················· | ····················· | ···················· | ···················· | ···················· |
| H | 693 hCoV-19/Canada/QC-LSPQ-L00231139/2020\|EPI_ISL_5359 | ···················· | ···················· | ····················· | ···················· | ···················· | ···················· |
| H | 694 hCoV-19/Canada/QC-LSPQ-L00231156/2020\|EPI_ISL_5359 | ···················· | ···················· | ····················· | ···················· | ···················· | ···················· |
| H | 695 hCoV-19/Canada/QC-LSPQ-L00231158/2020\|EPI_ISL_5359 | ···················· | ···················· | ····················· | ···················· | ···················· | ···················· |
| H | 696 hCoV-19/Canada/QC-LSPQ-L00231230/2020\|EPI_ISL_5359 | ···················· | ···················· | ····················· | ···················· | ···················· | ···················· |
| H | 697 hCoV-19/Canada/QC-LSPQ-L00231249/2020\|EPI_ISL_5359 | ···················· | ···················· | ····················· | ···················· | ···················· | ···················· |
| H | 698 hCoV-19/Canada/QC-LSPQ-L00231271/2020\|EPI_ISL_5359 | ···················· | ···················· | ····················· | ···················· | ···················· | ···················· |
| H | 699 hCoV-19/Canada/QC-LSPQ-L00231277/2020\|EPI_ISL_5359 | ···················· | ···················· | ····················· | ···················· | ···················· | ···················· |
| H | 700 hCoV-19/Canada/QC-LSPQ-L00231296/2020\|EPI_ISL_5359 | ···················· | ···················· | ····················· | ···················· | ···················· | ···················· |
| H | 701 hCoV-19/Canada/QC-LSPQ-L00231304/2020\|EPI_ISL_5359 | ···················· | ···················· | ····················· | ···················· | ···················· | ···················· |
| H | 702 hCoV-19/Canada/QC-LSPQ-L00231415/2020\|EPI_ISL_5359 | ···················· | ···················· | ····················· | ···················· | ···················· | ···················· |
| H | 703 hCoV-19/Canada/QC-LSPQ-L00231416/2020\|EPI_ISL_5359 | ···················· | ···················· | ····················· | ···················· | ···················· | ···················· |
| H | 704 hCoV-19/Canada/QC-LSPQ-L00231456/2020\|EPI_ISL_5359 | ···················· | ···················· | ····················· | ···················· | ···················· | ···················· |
| H | 705 hCoV-19/Canada/QC-LSPQ-L00231569/2020\|EPI_ISL_5359 | ···················· | ···················· | ····················· | ···················· | ···················· | ···················· |
| H | 706 hCoV-19/Canada/QC-LSPQ-L00231634/2020\|EPI_ISL_5359 | ···················· | ···················· | ····················· | ···················· | ···················· | ···················· |
| H | 707 hCoV-19/Canada/QC-LSPQ-L00231779/2020\|EPI_ISL_5359 | ···················· | ···················· | ····················· | ···················· | ···················· | ···················· |
| H | 708 hCoV-19/Canada/QC-LSPQ-L00231805/2020\|EPI_ISL_5359 | ···T················ | ···················· | ····················· | ···················· | ···················· | ···················· |
| H | 709 hCoV-19/Canada/QC-LSPQ-L00231814/2020\|EPI_ISL_5359 | ···················· | ···················· | ····················· | ···················· | ···················· | ···················· |
| H | 710 hCoV-19/Canada/QC-LSPQ-L00231860/2020\|EPI_ISL_5359 | ···················· | ···················· | ····················· | ···················· | ···················· | ···················· |
| H | 711 hCoV-19/Canada/QC-LSPQ-L00231946/2020\|EPI_ISL_5359 | ···················· | ···················· | ····················· | ···················· | ···················· | ···················· |
| H | 712 hCoV-19/Canada/QC-LSPQ-L00231966/2020\|EPI_ISL_5359 | ···················· | ···················· | ····················· | ···················· | ···················· | ···················· |
| H | 713 hCoV-19/Canada/QC-LSPQ-L00232004/2020\|EPI_ISL_5359 | ···················· | ···················· | ····················· | ···················· | ···················· | ···················· |
| H | 714 hCoV-19/Canada/QC-LSPQ-L00232064/2020\|EPI_ISL_5359 | ···················· | ···················· | ····················· | ···················· | ···················· | ···················· |
| H | 715 hCoV-19/Canada/QC-LSPQ-L00232070/2020\|EPI_ISL_5359 | ···················· | ···················· | ····················· | ···················· | ···················· | ···················· |
| H | 716 hCoV-19/Canada/QC-LSPQ-L00232073/2020\|EPI_ISL_5359 | ···················· | ···················· | ····················· | ···················· | ···················· | ···················· |
| H | 717 hCoV-19/Canada/QC-LSPQ-L00232080/2020\|EPI_ISL_5359 | ···················· | ···················· | ····················· | ···················· | ···················· | ···················· |
| H | 718 hCoV-19/Canada/QC-LSPQ-L00232097/2020\|EPI_ISL_5359 | ···················· | ···················· | ····················· | ···················· | ···················· | ···················· |
| H | 719 hCoV-19/Canada/QC-LSPQ-L00232098/2020\|EPI_ISL_5359 | ···················· | ···················· | ····················· | ···················· | ···················· | ···················· |
| H | 720 hCoV-19/Canada/QC-LSPQ-L00232101/2020\|EPI_ISL_5359 | ···················· | ···················· | ····················· | ···················· | ···················· | ···················· |
| H | 721 hCoV-19/Canada/QC-LSPQ-L00232107/2020\|EPI_ISL_5359 | ···················· | ···················· | ····················· | ···················· | ···················· | ···················· |
| H | 722 hCoV-19/Canada/QC-LSPQ-L00232113/2020\|EPI_ISL_5359 | ···················· | ···················· | ····················· | ···················· | ···················· | ···················· |
| H | 723 hCoV-19/Canada/QC-LSPQ-L00232148/2020\|EPI_ISL_5359 | ···················· | ···················· | ····················· | ···················· | ···················· | ···················· |
| H | 724 hCoV-19/Canada/QC-LSPQ-L00232187/2020\|EPI_ISL_5359 | ···················· | ···················· | ····················· | ···················· | ···················· | ···················· |
| H | 725 hCoV-19/Canada/QC-LSPQ-L00232239/2020\|EPI_ISL_5359 | ···················· | ···················· | ····················· | ···················· | ···················· | ···················· |
| H | 726 hCoV-19/Canada/QC-LSPQ-L00232271/2020\|EPI_ISL_5359 | ···················· | ···················· | ····················· | ···················· | ···················· | ···················· |
| H | 727 hCoV-19/Canada/QC-LSPQ-L00232439/2020\|EPI_ISL_5359 | ···················· | ···················· | ····················· | ···················· | ···················· | ···················· |
| H | 728 hCoV-19/Canada/QC-LSPQ-L00232447/2020\|EPI_ISL_5359 | ···················· | ·A·················· | ····················· | ···················· | ···················· | ···················· |
| H | 729 hCoV-19/England/QEUH-961DEF/2020\|EPI_ISL_537030\|20 | ···················· | ···················· | ····················· | ···················· | ···················· | ···················· |
| H | 730 hCoV-19/England/QEUH-961C5B/2020\|EPI_ISL_537031\|20 | ···················· | ···················· | ····················· | ···················· | ···················· | ···················· |
| H | 731 hCoV-19/England/QEUH-961D94/2020\|EPI_ISL_537032\|20 | ···················· | ·A·················· | ····················· | ···················· | ···················· | ···A················ |
| H | 732 hCoV-19/England/QEUH-961620/2020\|EPI_ISL_537033\|20 | ···················· | ·A·················· | ····················· | ···················· | ···················· | ···················· |
| H | 733 hCoV-19/England/QEUH-961AF7/2020\|EPI_ISL_537034\|20 | ···················· | ···················· | ····················· | ···················· | ···················· | ···················· |
| H | 734 hCoV-19/England/QEUH-95DE6B/2020\|EPI_ISL_537036\|20 | ···················· | ···················· | ····················· | ···················· | ···················· | ···················· |
| H | 735 hCoV-19/England/QEUH-95E107/2020\|EPI_ISL_537037\|20 | ···················· | ···················· | ····················· | ···················· | ···················· | ···················· |
| H | 736 hCoV-19/England/QEUH-96168A/2020\|EPI_ISL_537038\|20 | ···················· | ···················· | ····················· | ···················· | ···················· | ···················· |
| H | 737 hCoV-19/England/QEUH-961A8E/2020\|EPI_ISL_537039\|20 | ···················· | ···················· | ····················· | ···················· | ···················· | ···················· |
| H | 738 hCoV-19/Canada/ON-OICR-SLB2836/2020\|EPI_ISL_538360 | ···················· | ···················· | ····················· | ···················· | ···················· | ···················· |
| H | 739 hCoV-19/Canada/ON-OICR-SLB2838/2020\|EPI_ISL_538361 | ···················· | ···················· | ····················· | ···················· | ···················· | ···················· |
| H | 740 hCoV-19/Canada/ON-OICR-SLB2841/2020\|EPI_ISL_538362 | ···················· | ···················· | ····················· | ···················· | ···················· | ···················· |
| H | 741 hCoV-19/Canada/ON-OICR-SLB2842/2020\|EPI_ISL_538363 | ···················· | ···················· | ····················· | ···················· | ···················· | ···················· |
| H | 742 hCoV-19/Canada/ON-OICR-SLB2843/2020\|EPI_ISL_538364 | ···················· | ···················· | ····················· | ···················· | ···················· | ···················· |
| H | 743 hCoV-19/Canada/ON-OICR-SLB2844/2020\|EPI_ISL_538365 | ···················· | ···················· | ····················· | ···················· | ···················· | ···················· |
| H | 744 hCoV-19/Canada/ON-OICR-SLB2847/2020\|EPI_ISL_538367 | ···················· | ···················· | ····················· | ···················· | ···················· | ···················· |
| H | 745 hCoV-19/Canada/ON-OICR-SLB2848/2020\|EPI_ISL_538368 | ···················· | ···················· | ····················· | ···················· | ···················· | ···················· |
| H | 746 hCoV-19/Canada/ON-OICR-SLB2849/2020\|EPI_ISL_538369 | ···················· | ···················· | ····················· | ···················· | ···················· | ···················· |
| H | 747 hCoV-19/India/TG-CCMB-AA403/2020\|EPI_ISL_539690\|20 | ···················· | ···················· | ····················· | ···················· | ···················· | ···················· |
| H | 748 hCoV-19/India/TG-CCMB-AB927/2020\|EPI_ISL_539691\|20 | ···················· | ···················· | ····················· | ···················· | ···················· | ···················· |
| H | 749 hCoV-19/India/TG-CCMB-AA373/2020\|EPI_ISL_539692\|20 | ···················· | ···················· | ····················· | ···················· | ···················· | ···················· |
| H | 750 hCoV-19/India/TG-CCMB-AA770/2020\|EPI_ISL_539693\|20 | ···················· | ···················· | ····················· | ···················· | ···················· | ···················· |
| H | 751 hCoV-19/India/TG-CCMB-AA456/2020\|EPI_ISL_539694\|20 | ···················· | ···················· | ····················· | ···················· | ···················· | ···················· |
| H | 752 hCoV-19/India/TG-CCMB-AA867/2020\|EPI_ISL_539695\|20 | ···················· | ···················· | ····················· | ···················· | ···················· | ···················· |
| H | 753 hCoV-19/India/TG-CCMB-AA868/2020\|EPI_ISL_539696\|20 | ···················· | ···················· | ····················· | ···················· | ···················· | ···················· |
| H | 754 hCoV-19/India/TG-CCMB-AA869/2020\|EPI_ISL_539697\|20 | ···················· | ···················· | ····················· | ···················· | ···················· | ···················· |
| H | 755 hCoV-19/India/TG-CCMB-AA870/2020\|EPI_ISL_539698\|20 | ···················· | ···················· | ····················· | ···················· | ···················· | ···················· |
| H | 756 hCoV-19/India/TG-CCMB-AA789/2020\|EPI_ISL_539699\|20 | ···················· | ···················· | ····················· | ···················· | ···················· | ···················· |
| H | 757 hCoV-19/England/MILK-99236B/2020\|EPI_ISL_540674\|20 | ···················· | ···················· | ····················· | ···················· | ···················· | ···················· |
| H | 758 hCoV-19/England/MILK-992477/2020\|EPI_ISL_540682\|20 | ···················· | ···················· | ····················· | ···················· | ···················· | ···················· |
| H | 759 hCoV-19/England/MILK-992486/2020\|EPI_ISL_540683\|20 | ···················· | ···················· | ····················· | ···················· | ···················· | ···················· |

| | | | | | | | | |
|---|---|---|---|---|---|---|---|---|
| H | 760 hCoV-19/England/QEUH-998FAD/2020\|EPI_ISL_541800\|20 | .................... | ................... | .................... | .................... | ................... | .................... | |
| H | 761 hCoV-19/England/QEUH-999452/2020\|EPI_ISL_541801\|20 | .................... | ................... | .................... | .................... | ................... | .................... | |
| H | 762 hCoV-19/Scotland/QEUH-99909A/2020\|EPI_ISL_541802\|2 | .................... | ................... | .................... | .................... | ................... | .................... | |
| H | 763 hCoV-19/England/QEUH-99908B/2020\|EPI_ISL_541803\|20 | .................... | ................... | .................... | .................... | ................... | .................... | |
| H | 764 hCoV-19/England/QEUH-998E73/2020\|EPI_ISL_541804\|20 | .................... | ................... | .................... | .................... | .....T............. | .................... | |
| H | 765 hCoV-19/England/QEUH-99922B/2020\|EPI_ISL_541805\|20 | .................... | ................... | .................... | .................... | ................... | .................... | |
| H | 766 hCoV-19/Northern Ireland/QEUH-998EEC/2020\|EPI_ISL_ | .................... | ................... | .................... | .................... | ................... | .................... | |
| H | 767 hCoV-19/England/QEUH-999391/2020\|EPI_ISL_541808\|20 | .................... | ................... | .................... | .................... | ................... | .................... | |
| H | 768 hCoV-19/Scotland/QEUH-99916A/2020\|EPI_ISL_541809\|2 | .................... | ................... | .................... | .................... | ................... | .................... | |
| H | 769 hCoV-19/England/QEUH-960D1D/2020\|EPI_ISL_532001\|20 | .................... | ................... | .................... | .................... | ................... | .................... | |
| H | 770 hCoV-19/Scotland/QEUH-960D2C/2020\|EPI_ISL_532003\|2 | .................... | ................... | .................... | .................... | ................... | .................... | |
| H | 771 hCoV-19/Scotland/QEUH-960F44/2020\|EPI_ISL_532004\|2 | .................... | ................... | .................... | .................... | ................... | .................... | |
| H | 772 hCoV-19/Scotland/QEUH-960797/2020\|EPI_ISL_532005\|2 | .................... | ................... | .................... | .................... | ................... | .................... | |
| H | 773 hCoV-19/England/QEUH-960885/2020\|EPI_ISL_532006\|20 | .................... | ................... | .................... | .................... | ................... | .................... | |
| H | 774 hCoV-19/England/QEUH-960858/2020\|EPI_ISL_532008\|20 | .................... | ................... | .................... | .................... | ................... | .................... | |
| H | 775 hCoV-19/England/QEUH-9612B3/2020\|EPI_ISL_532009\|20 | .................... | ................... | .................... | .................... | ................... | .................... | |
| H | 776 hCoV-19/England/QEUH-960A61/2020\|EPI_ISL_532010\|20 | .................... | ................... | .................... | .................... | ................... | .................... | |
| H | 777 hCoV-19/England/QEUH-960296/2020\|EPI_ISL_532011\|20 | .................... | ................... | .................... | .................... | ................... | .................... | |
| H | 778 hCoV-19/England/QEUH-9604DC/2020\|EPI_ISL_532014\|20 | .................... | ................... | .................... | .................... | ................... | .................... | |
| H | 779 hCoV-19/England/QEUH-960612/2020\|EPI_ISL_532015\|20 | .................... | ................... | .................... | .................... | ................... | .................... | |
| H | 780 hCoV-19/Northern Ireland/QEUH-960E56/2020\|EPI_ISL_ | .................... | ................... | .................... | .................... | ................... | .................... | |
| H | 781 hCoV-19/Scotland/QEUH-96109B/2020\|EPI_ISL_532018\|2 | .................... | ................... | .................... | .................... | ................... | .................... | |
| H | 782 hCoV-19/Northern Ireland/QEUH-960DD1/2020\|EPI_ISL_ | .................... | ................... | .................... | .................... | ................... | .................... | |
| H | 784 hCoV-19/England/QEUH-960A8F/2020\|EPI_ISL_532021\|20 | .................... | ................... | .................... | .................... | ................... | .................... | |
| H | 785 hCoV-19/England/QEUH-9602F0/2020\|EPI_ISL_532022\|20 | .................... | ................... | .................... | .................... | ................... | .................... | |
| H | 786 hCoV-19/England/QEUH-960E74/2020\|EPI_ISL_532023\|20 | .................... | ................... | .................... | .................... | ................... | .................... | |
| H | 787 hCoV-19/Scotland/QEUH-961004/2020\|EPI_ISL_532024\|2 | .................... | .A................. | .................... | .................... | ................... | .................... | |
| H | 788 hCoV-19/England/QEUH-961295/2020\|EPI_ISL_532025\|20 | .................... | ................... | .................... | .................... | ................... | .................... | |
| H | 789 hCoV-19/England/QEUH-9605AC/2020\|EPI_ISL_532026\|20 | .................... | ................... | .................... | .................... | ................... | .................... | |
| H | 790 hCoV-19/England/QEUH-960919/2020\|EPI_ISL_532027\|20 | .................... | ................... | .................... | .................... | ................... | .................... | |
| H | 791 hCoV-19/England/QEUH-5E8CD8/2020\|EPI_ISL_533322\|20 | .................... | ................... | .................... | .................... | ................... | .................... | |
| H | 792 hCoV-19/England/QEUH-5E9114/2020\|EPI_ISL_533324\|20 | .................... | ................... | .................... | .................... | ................... | .................... | |
| H | 793 hCoV-19/England/QEUH-5DF339/2020\|EPI_ISL_533325\|20 | .................... | ................... | .................... | .................... | ................... | .................... | |
| H | 794 hCoV-19/England/QEUH-5E5D32/2020\|EPI_ISL_533326\|20 | .................... | ................... | .................... | .................... | ................... | .................... | |
| H | 795 hCoV-19/England/QEUH-5E2CA1/2020\|EPI_ISL_533329\|20 | .................... | ................... | .................... | .................... | ................... | .................... | |
| H | 796 hCoV-19/England/QEUH-5EB6B9/2020\|EPI_ISL_533331\|20 | .................... | ................... | .................... | .................... | ................... | .................... | |
| H | 797 hCoV-19/England/QEUH-5DFB31/2020\|EPI_ISL_533332\|20 | .................... | ................... | .................... | .................... | ................... | .................... | |
| H | 798 hCoV-19/England/QEUH-5DCF38/2020\|EPI_ISL_533333\|20 | .................... | ................... | .................... | .................... | ................... | .................... | |
| H | 799 hCoV-19/England/QEUH-5DA2C8/2020\|EPI_ISL_533335\|20 | .................... | ................... | .................... | .................... | ................... | .................... | |
| H | 800 hCoV-19/England/QEUH-5E03BD/2020\|EPI_ISL_533336\|20 | .................... | ................... | .................... | .................... | ................... | .................... | |
| H | 801 hCoV-19/England/QEUH-5EA5EA/2020\|EPI_ISL_533337\|20 | .................... | ................... | .................... | .................... | ................... | .................... | |
| H | 802 hCoV-19/England/QEUH-5E05D5/2020\|EPI_ISL_533338\|20 | .................... | ................... | .................... | .................... | ................... | .................... | |
| H | 803 hCoV-19/England/QEUH-5DEA44/2020\|EPI_ISL_533339\|20 | .................... | ................... | .................... | .................... | ................... | .................... | |
| H | 804 hCoV-19/England/QEUH-5EA18C/2020\|EPI_ISL_533341\|20 | .................... | ................... | .................... | .................... | ................... | .................... | |
| H | 805 hCoV-19/England/QEUH-5DA170/2020\|EPI_ISL_533342\|20 | .................... | ................... | .................... | .................... | ................... | .................... | |
| H | 806 hCoV-19/Scotland/QEUH-5E0F31/2020\|EPI_ISL_533343\|2 | .................... | ................... | .................... | .................... | ................... | .................... | |
| H | 807 hCoV-19/England/QEUH-5DF7B5/2020\|EPI_ISL_533344\|20 | .................... | ................... | .................... | .................... | ................... | .................... | |
| H | 808 hCoV-19/England/QEUH-544D9B/2020\|EPI_ISL_533345\|20 | .................... | ................... | .................... | .................... | ................... | .................... | |
| H | 809 hCoV-19/Scotland/QEUH-545EF1/2020\|EPI_ISL_533346\|2 | .................... | .A................. | .................... | .................... | ................... | .................... | |
| H | 810 hCoV-19/England/QEUH-546DD5/2020\|EPI_ISL_533347\|20 | .................... | ................... | .................... | .................... | ................... | .................... | |
| H | 811 hCoV-19/England/QEUH-543BDE/2020\|EPI_ISL_533348\|20 | .................... | ................... | .................... | .................... | ................... | .................... | |
| H | 812 hCoV-19/England/QEUH-542D9D/2020\|EPI_ISL_533349\|20 | .................... | ................... | .................... | .................... | ................... | .................... | |
| H | 813 hCoV-19/Scotland/QEUH-5443D5/2020\|EPI_ISL_533350\|2 | .................... | ..................A | .................... | .................... | ................... | .................... | 1 |
| H | 814 hCoV-19/Scotland/QEUH-546081/2020\|EPI_ISL_533351\|2 | .................... | ................... | .................... | .................... | ................... | .................... | |
| H | 815 hCoV-19/England/QEUH-548939/2020\|EPI_ISL_533352\|20 | .................... | ................... | .................... | .................... | ................... | .................... | |
| H | 816 hCoV-19/England/QEUH-545BAF/2020\|EPI_ISL_533353\|20 | .................... | ................... | .................... | .................... | ................... | .................... | |
| H | 817 hCoV-19/England/QEUH-543190/2020\|EPI_ISL_533356\|20 | .................... | ................... | .................... | .................... | ................... | .................... | |
| H | 818 hCoV-19/Wales/QEUH-5458A8/2020\|EPI_ISL_533357\|2020 | .................... | ................... | .................... | .................... | ................... | .................... | |
| H | 819 hCoV-19/Scotland/QEUH-96BBFA/2020\|EPI_ISL_534653\|2 | .................... | ................... | .................... | .................... | ................... | .................... | |
| H | 820 hCoV-19/Scotland/QEUH-96B3C5/2020\|EPI_ISL_534656\|2 | .................... | ................... | .................... | .................... | ................... | .................... | |
| H | 821 hCoV-19/Scotland/QEUH-96B547/2020\|EPI_ISL_534657\|2 | .................... | ................... | .................... | .................... | ................... | .................... | |
| H | 822 hCoV-19/Scotland/QEUH-96B42C/2020\|EPI_ISL_534658\|2 | .................... | ................... | .................... | .................... | ................... | .................... | |
| H | 823 hCoV-19/Scotland/QEUH-96B495/2020\|EPI_ISL_534659\|2 | .................... | ................... | .................... | .................... | ................... | .................... | |
| H | 824 hCoV-19/Scotland/QEUH-96B5DE/2020\|EPI_ISL_534661\|2 | .................... | ................... | .................... | .................... | ................... | .................... | |
| H | 825 hCoV-19/Scotland/QEUH-96B77E/2020\|EPI_ISL_534662\|2 | .................... | ................... | .................... | .................... | ................... | .................... | |
| H | 826 hCoV-19/Scotland/QEUH-96B51A/2020\|EPI_ISL_534663\|2 | .................... | ................... | .................... | .................... | ................... | .................... | |
| H | 827 hCoV-19/Scotland/QEUH-96B0DD/2020\|EPI_ISL_534664\|2 | .................... | ................... | .................... | .................... | ................... | .................... | |
| H | 828 hCoV-19/Scotland/QEUH-96A96A/2020\|EPI_ISL_534665\|2 | .................... | ................... | .................... | .................... | ................... | .................... | |
| H | 829 hCoV-19/Scotland/QEUH-96B574/2020\|EPI_ISL_534666\|2 | .................... | .A................. | .................... | .................... | ................... | ..A................. | |
| H | 830 hCoV-19/Scotland/QEUH-96B644/2020\|EPI_ISL_534667\|2 | .................... | ................... | .................... | .................... | ................... | .................... | |
| H | 831 hCoV-19/Scotland/QEUH-96B0EC/2020\|EPI_ISL_534668\|2 | .................... | ................... | .................... | .................... | ................... | .................... | |
| H | 832 hCoV-19/Scotland/QEUH-96B3B6/2020\|EPI_ISL_534669\|2 | .................... | ................... | .................... | .................... | ................... | .................... | |
| H | 833 hCoV-19/Scotland/QEUH-96BEC4/2020\|EPI_ISL_534670\|2 | .................... | ................... | .................... | .................... | ................... | .................... | |
| H | 834 hCoV-19/Scotland/QEUH-96B2D7/2020\|EPI_ISL_534671\|2 | .................... | ................... | .................... | .................... | .....T............. | .................... | |
| H | 835 hCoV-19/Scotland/QEUH-96AAD0/2020\|EPI_ISL_534672\|2 | .................... | ................... | .................... | .................... | ................... | .................... | |
| H | 836 hCoV-19/Scotland/QEUH-96BF49/2020\|EPI_ISL_534673\|2 | .................... | ................... | .................... | .................... | ................... | .................... | |
| H | 837 hCoV-19/Scotland/QEUH-96AAC1/2020\|EPI_ISL_534674\|2 | .................... | ................... | .................... | .................... | ................... | .................... | |
| H | 838 hCoV-19/Scotland/QEUH-96B76F/2020\|EPI_ISL_534675\|2 | .................... | ................... | .................... | .................... | ................... | .................... | |
| H | 839 hCoV-19/Scotland/QEUH-96BD6D/2020\|EPI_ISL_534676\|2 | .................... | .A................. | .................... | .................... | ................... | .................... | |
| H | 840 hCoV-19/Scotland/QEUH-96B6DB/2020\|EPI_ISL_534677\|2 | .................... | ................... | .................... | .................... | ................... | .................... | |

| | | | | | | | |
|---|---|---|---|---|---|---|---|
| H | 841 hCoV-19/Scotland/QEUH-96BA0C/2020\|EPI_ISL_534678\|2 | ···················· | ·A·················· | ···················· | ···················· | ···················· | ···················· |
| H | 842 hCoV-19/Scotland/QEUH-96B635/2020\|EPI_ISL_534679\|2 | ···················· | ···················· | ···················· | ···················· | ···················· | ···················· |
| H | 843 hCoV-19/Scotland/QEUH-96A9E2/2020\|EPI_ISL_534680\|2 | ···················· | ···················· | ···················· | ···················· | ···················· | ···················· |
| H | 844 hCoV-19/Scotland/QEUH-96B9A5/2020\|EPI_ISL_534681\|2 | ···················· | ···················· | ···················· | ···················· | ···················· | ···················· |
| H | 845 hCoV-19/Scotland/QEUH-96BF2B/2020\|EPI_ISL_534683\|2 | ···················· | ···················· | ···················· | ···················· | ···················· | ···················· |
| H | 846 hCoV-19/Scotland/QEUH-96ACCB/2020\|EPI_ISL_534684\|2 | ···················· | ···················· | ···················· | ···················· | ···················· | ···················· |
| H | 847 hCoV-19/Scotland/QEUH-96ADE6/2020\|EPI_ISL_534685\|2 | ···················· | ···················· | ···················· | ···················· | ···················· | ···················· |
| H | 848 hCoV-19/Scotland/QEUH-96B204/2020\|EPI_ISL_534686\|2 | ···················· | ···················· | ···················· | ···················· | ···················· | ···················· |
| H | 849 hCoV-19/Scotland/QEUH-96B6CC/2020\|EPI_ISL_534687\|2 | ···················· | ···················· | ···················· | ···················· | ···················· | ···················· |
| H | 850 hCoV-19/Scotland/QEUH-96B37A/2020\|EPI_ISL_534688\|2 | ···················· | ···················· | ···················· | ···················· | ···················· | ···················· |
| H | 851 hCoV-19/USA/MD-MDH-0140/2020\|EPI_ISL_534700\|2020-0 | ···················· | ············C······· | ···················· | ···················· | ···················· | ···················· |
| H | 852 hCoV-19/USA/MD-MDH-0141/2020\|EPI_ISL_534701\|2020-0 | ···················· | ············C······· | ···················· | ···················· | ···················· | ···················· |
| H | 853 hCoV-19/USA/MD-MDH-0142/2020\|EPI_ISL_534702\|2020-0 | ···················· | ···················· | ···················· | ···················· | ···················· | ···················· |
| H | 854 hCoV-19/USA/MD-MDH-0143/2020\|EPI_ISL_534703\|2020-0 | ···················· | ···················· | ···················· | ···················· | ···················· | ···················· |
| H | 855 hCoV-19/USA/MD-MDH-0165/2020\|EPI_ISL_534704\|2020-0 | ···················· | ···················· | ···················· | ···················· | ···················· | ···················· |
| H | 856 hCoV-19/USA/MD-MDH-0166/2020\|EPI_ISL_534705\|2020-0 | ···················· | ···················· | ···················· | ···················· | ···················· | ···················· |
| H | 857 hCoV-19/USA/MD-MDH-0169/2020\|EPI_ISL_534706\|2020-0 | ···················· | ···················· | ···················· | ···················· | ···················· | ···················· |
| H | 858 hCoV-19/USA/MD-MDH-0171/2020\|EPI_ISL_534708\|2020-0 | ···················· | ···················· | ···················· | ···················· | ···················· | ···················· |
| H | 859 hCoV-19/USA/MD-MDH-0172/2020\|EPI_ISL_534709\|2020-0 | ···················· | ···················· | ···················· | ···················· | ···················· | ···················· |
| H | 860 hCoV-19/USA/MD-MDH-0173/2020\|EPI_ISL_534710\|2020-0 | ···················· | ···················· | ···················· | ···················· | ···················· | ···················· |
| H | 861 hCoV-19/USA/MD-MDH-0174/2020\|EPI_ISL_534711\|2020-0 | ···················· | ···················· | ···················· | ···················· | ···················· | ···················· |
| H | 862 hCoV-19/USA/MD-MDH-0176/2020\|EPI_ISL_534713\|2020-0 | ···················· | ···················· | ···················· | ···················· | ···················· | ···················· |
| H | 863 hCoV-19/USA/MD-MDH-0179/2020\|EPI_ISL_534716\|2020-0 | ···················· | ···················· | ···················· | ···················· | ···················· | ···················· |
| H | 864 hCoV-19/England/202390165/2020\|EPI_ISL_534717\|2020 | ···················· | ···················· | ···················· | ···················· | ···················· | ···················· |
| H | 865 hCoV-19/Canada/QC-LSPQ-L00232310/2020\|EPI_ISL_5359 | ···················· | ···················· | ···················· | ···················· | ···················· | ···················· |
| H | 866 hCoV-19/Canada/QC-LSPQ-L00232405/2020\|EPI_ISL_5359 | ···················· | ·A·················· | ···················· | ···················· | ···················· | ···················· |
| H | 867 hCoV-19/Canada/QC-LSPQ-L00232511/2020\|EPI_ISL_5359 | ···················· | ···················· | ···················· | ···················· | ···················· | ···················· |
| H | 868 hCoV-19/Canada/QC-LSPQ-L00232601/2020\|EPI_ISL_5359 | ···················· | ···················· | ···················· | ···················· | ···················· | ···················· |
| H | 869 hCoV-19/Canada/QC-LSPQ-L00232603/2020\|EPI_ISL_5359 | ···················· | ···················· | ···················· | ···················· | ···················· | ···················· |
| H | 870 hCoV-19/Canada/QC-LSPQ-L00232609/2020\|EPI_ISL_5359 | ···················· | ···················· | ···················· | ···················· | ···················· | ···················· |
| H | 871 hCoV-19/Canada/QC-LSPQ-L00232614/2020\|EPI_ISL_5359 | ···················· | ···················· | ···················· | ···················· | ···················· | ···················· |
| H | 872 hCoV-19/Canada/QC-LSPQ-L00232723/2020\|EPI_ISL_5359 | ···················· | ···················· | ···················· | ···················· | ···················· | ···················· |
| H | 873 hCoV-19/Canada/QC-LSPQ-L00232726/2020\|EPI_ISL_5359 | ···················· | ···················· | ···················· | ···················· | ···················· | ···················· |
| H | 874 hCoV-19/Canada/QC-LSPQ-L00232810/2020\|EPI_ISL_5359 | ···················· | ···················· | ···················· | ···················· | ···················· | ···················· |
| H | 875 hCoV-19/Canada/QC-LSPQ-L00232813/2020\|EPI_ISL_5359 | ···················· | ···················· | ···················· | ···················· | ···················· | ···················· |
| H | 876 hCoV-19/Canada/QC-LSPQ-L00232822/2020\|EPI_ISL_5359 | ···················· | ···················· | ···················· | ···················· | ···················· | ···················· |
| H | 877 hCoV-19/Canada/QC-LSPQ-L00232866/2020\|EPI_ISL_5359 | ···················· | ···················· | ···················· | ···················· | ···················· | ···················· |
| H | 878 hCoV-19/USA/MD-MDH-0181/2020\|EPI_ISL_538265\|2020-0 | ···················· | ···················· | ···················· | ···················· | ···················· | ···················· |
| H | 879 hCoV-19/USA/MD-MDH-0182/2020\|EPI_ISL_538266\|2020-0 | ···················· | ···················· | ···················· | ···················· | ···················· | ···················· |
| H | 880 hCoV-19/USA/MD-MDH-0183/2020\|EPI_ISL_538267\|2020-0 | ···················· | ···················· | ···················· | ···················· | ···················· | ···················· |
| H | 881 hCoV-19/USA/MD-MDH-0184/2020\|EPI_ISL_538268\|2020-0 | ···················· | ···················· | ···················· | ···················· | ···················· | ···················· |
| H | 882 hCoV-19/USA/MD-MDH-0185/2020\|EPI_ISL_538269\|2020-0 | ···················· | ···················· | ···················· | ···················· | ···················· | ···················· |
| H | 883 hCoV-19/USA/MD-MDH-0186/2020\|EPI_ISL_538270\|2020-0 | ···················· | ···················· | ···················· | ···················· | ···················· | ···················· |
| H | 884 hCoV-19/USA/MD-MDH-0187/2020\|EPI_ISL_538271\|2020-0 | ···················· | ···················· | ···················· | ···················· | ···················· | ···················· |
| H | 885 hCoV-19/USA/MD-MDH-0189/2020\|EPI_ISL_538272\|2020-0 | ···················· | ···················· | ···················· | ···················· | ···················· | ···················· |
| H | 886 hCoV-19/USA/MD-MDH-0190/2020\|EPI_ISL_538273\|2020-0 | ···················· | ···················· | ···················· | ···················· | ···················· | ···················· |
| H | 887 hCoV-19/USA/MD-MDH-0191/2020\|EPI_ISL_538274\|2020-0 | ···················· | ···················· | ···················· | ···················· | ···················· | ···················· |
| H | 888 hCoV-19/USA/TX-DSHS-0342/2020\|EPI_ISL_538280\|2020- | ···················· | ···················· | ···················· | ···················· | ···················· | ···················· |
| H | 889 hCoV-19/Spain/AR-ISCIII-201435/2020\|EPI_ISL_539500 | ···················· | ···················· | ···················· | ···················· | ···················· | ···················· |
| H | 890 hCoV-19/Spain/AR-ISCIII-201587/2020\|EPI_ISL_539501 | ···················· | ···················· | ···················· | ···················· | ···················· | ···················· |
| H | 891 hCoV-19/Spain/AR-ISCIII-201588/2020\|EPI_ISL_539502 | ···················· | ···················· | ···················· | ···················· | ···················· | ···················· |
| H | 892 hCoV-19/Spain/AR-ISCIII-201596/2020\|EPI_ISL_539503 | ···················· | ···················· | ···················· | ···················· | ···················· | ···················· |
| H | 893 hCoV-19/Spain/AR-ISCIII-201597/2020\|EPI_ISL_539504 | ···················· | ···················· | ···················· | ···················· | ···················· | ···················· |
| H | 894 hCoV-19/Spain/AR-ISCIII-201664/2020\|EPI_ISL_539505 | ···················· | ···················· | ···················· | ···················· | ···················· | ···················· |
| H | 895 hCoV-19/Spain/AR-ISCIII-201665/2020\|EPI_ISL_539506 | ···················· | ···················· | ···················· | ···················· | ···················· | ···················· |
| H | 896 hCoV-19/Spain/AR-ISCIII-201672/2020\|EPI_ISL_539507 | ···················· | ···················· | ···················· | ···················· | ···················· | ···················· |
| H | 897 hCoV-19/Spain/AR-ISCIII-201673/2020\|EPI_ISL_539508 | ···················· | ···················· | ···················· | ···················· | ···················· | ···················· |
| H | 898 hCoV-19/Spain/AR-ISCIII-201687/2020\|EPI_ISL_539509 | ···················· | ···················· | ···················· | ···················· | ···················· | ···················· |
| H | 899 hCoV-19/Spain/AR-ISCIII-201688/2020\|EPI_ISL_539510 | ···················· | ···················· | ···················· | ···················· | ···················· | ···················· |
| H | 900 hCoV-19/Spain/AR-ISCIII-201692/2020\|EPI_ISL_539511 | ···················· | ···················· | ···················· | ···················· | ···················· | ···················· |
| H | 901 hCoV-19/Spain/AR-ISCIII-201693/2020\|EPI_ISL_539512 | ···················· | ···················· | ···················· | ···················· | ···················· | ···················· |
| H | 902 hCoV-19/Spain/AR-ISCIII-201779/2020\|EPI_ISL_539513 | ···················· | ···················· | ···················· | ···················· | ···················· | ···················· |
| H | 903 hCoV-19/Spain/AR-ISCIII-201780/2020\|EPI_ISL_539514 | ···················· | ···················· | ···················· | ···················· | ···················· | ···················· |
| H | 904 hCoV-19/Spain/AR-ISCIII-201782/2020\|EPI_ISL_539515 | ···················· | ···················· | ···················· | ···················· | ···················· | ···················· |
| H | 905 hCoV-19/Spain/AR-ISCIII-201784/2020\|EPI_ISL_539516 | ···················· | ···················· | ···················· | ···················· | ···················· | ···················· |
| H | 906 hCoV-19/Spain/AR-ISCIII-201848/2020\|EPI_ISL_539517 | ···················· | ···················· | ···················· | ···················· | ···················· | ···················· |
| H | 907 hCoV-19/Spain/AR-ISCIII-201849/2020\|EPI_ISL_539518 | ···················· | ···················· | ···················· | ···················· | ···················· | ···················· |
| H | 908 hCoV-19/Spain/AR-ISCIII-201850/2020\|EPI_ISL_539519 | ···················· | ···················· | ···················· | ···················· | ···················· | ···················· |
| H | 909 hCoV-19/Spain/CE-ISCIII-202293/2020\|EPI_ISL_539520 | ···················· | ···················· | ···················· | ···················· | ···················· | ···················· |
| H | 910 hCoV-19/Spain/CE-ISCIII-203352/2020\|EPI_ISL_539521 | ···················· | ···················· | ···················· | ···················· | ···················· | ···················· |
| H | 911 hCoV-19/Spain/CE-ISCIII-204881/2020\|EPI_ISL_539522 | ···················· | ···················· | ···················· | ···················· | ···················· | ···················· |
| H | 912 hCoV-19/Spain/CL-ISCIII-202362/2020\|EPI_ISL_539523 | ···················· | ···················· | ···················· | ···················· | ···················· | ···················· |
| H | 913 hCoV-19/Spain/CL-ISCIII-202468/2020\|EPI_ISL_539524 | ···················· | ···················· | ···················· | ···················· | ···················· | ···················· |
| H | 914 hCoV-19/Spain/CL-ISCIII-204996/2020\|EPI_ISL_539525 | ···················· | ···················· | ···················· | ···················· | ···················· | ···················· |
| H | 915 hCoV-19/Spain/CM-ISCIII-202018188/2020\|EPI_ISL_539 | ···················· | ···················· | ···················· | ···················· | ···················· | ···················· |
| H | 916 hCoV-19/Spain/CM-ISCIII-202018234/2020\|EPI_ISL_539 | ···················· | ···················· | ···················· | ···················· | ···················· | ···················· |
| H | 917 hCoV-19/Spain/CM-ISCIII-202018271/2020\|EPI_ISL_539 | ···················· | ···················· | ···················· | ···················· | ···················· | ·······G············ |
| H | 918 hCoV-19/Spain/CM-ISCIII-202018315/2020\|EPI_ISL_539 | ···················· | ···················· | ···················· | ···················· | ···················· | ···················· |
| H | 919 hCoV-19/Spain/CM-ISCIII-202018318/2020\|EPI_ISL_539 | ···················· | ···················· | ···················· | ···················· | ···················· | ···················· |
| H | 920 hCoV-19/Spain/CN-ISCIII-201048/2020\|EPI_ISL_539531 | ···················· | ···················· | ···················· | ···················· | ···················· | ···················· |

| | | | | | | | | |
|---|---|---|---|---|---|---|---|---|
| H | 921 | hCoV-19/Spain/CN-ISCIII-201810/2020\|EPI_ISL_539532 | ···················· | ···················· | ···················· | ···················· | ···················· | ···················· |
| H | 922 | hCoV-19/Spain/CT-ISCIII-2012563/2020\|EPI_ISL_53953 | ···················· | ···················· | ···················· | ···················· | ···················· | ···················· |
| H | 923 | hCoV-19/Spain/CT-ISCIII-2012564/2020\|EPI_ISL_53953 | ···················· | ···················· | ···················· | ···················· | ···················· | ···················· |
| H | 924 | hCoV-19/Spain/CT-ISCIII-2012566/2020\|EPI_ISL_53953 | ···················· | ···················· | ···················· | ···················· | ···················· | ···················· |
| H | 925 | hCoV-19/Spain/CT-ISCIII-2012568/2020\|EPI_ISL_53953 | ···················· | ···················· | ···················· | ···················· | ···················· | ···················· |
| H | 926 | hCoV-19/Spain/CT-ISCIII-2012571/2020\|EPI_ISL_53953 | ···················· | ···················· | ···················· | ···················· | ·······S············ | ···················· |
| H | 927 | hCoV-19/Spain/CT-ISCIII-2012572/2020\|EPI_ISL_53953 | ···················· | ···················· | ···················· | ···················· | ···················· | ···················· |
| H | 928 | hCoV-19/Spain/CT-ISCIII-2012573/2020\|EPI_ISL_53954 | ···················· | ···················· | ···················· | ···················· | ···················· | ···················· |
| H | 929 | hCoV-19/Spain/CT-ISCIII-2013585/2020\|EPI_ISL_53954 | ···················· | ···················· | ···················· | ···················· | ···················· | ···················· |
| H | 930 | hCoV-19/Spain/CT-ISCIII-2013586/2020\|EPI_ISL_53954 | ···················· | ···················· | ···················· | ···················· | ···················· | ···················· |
| H | 931 | hCoV-19/Spain/CT-ISCIII-2013591/2020\|EPI_ISL_53954 | ···················· | ···················· | ···················· | ···················· | ···················· | ···················· |
| H | 932 | hCoV-19/Spain/CT-ISCIII-2013592/2020\|EPI_ISL_53954 | ···················· | ···················· | ···················· | ···················· | ···················· | ···················· |
| H | 933 | hCoV-19/Spain/CT-ISCIII-2013597/2020\|EPI_ISL_53954 | ···················· | ···················· | ···················· | ···················· | ···················· | ···················· |
| H | 934 | hCoV-19/Spain/CT-ISCIII-202044/2020\|EPI_ISL_539549 | ···················· | ···················· | ···················· | ···················· | ···················· | ···················· |
| H | 935 | hCoV-19/Germany/NW-HHU-110/2020\|EPI_ISL_539587\|202 | ···················· | ···················· | ···················· | ···················· | ···················· | ···················· |
| H | 936 | hCoV-19/Germany/NW-HHU-114/2020\|EPI_ISL_539591\|202 | ···················· | ···················· | ···················· | ···················· | ···················· | ···················· |
| H | 937 | hCoV-19/Germany/NW-HHU-116/2020\|EPI_ISL_539593\|202 | ···················· | ···················· | ···················· | ···················· | ···················· | ···················· |
| H | 938 | hCoV-19/Germany/NW-HHU-80/2020\|EPI_ISL_539596\|2020 | ···················· | ···················· | ···················· | ···················· | ···················· | ···················· |
| H | 939 | hCoV-19/Germany/NW-HHU-83/2020\|EPI_ISL_539599\|2020 | ···················· | ···················· | ···················· | ···················· | ···················· | ···················· |
| H | 940 | hCoV-19/Belgium/ULG-10333/2020\|EPI_ISL_540500\|2020 | ···················· | ···················· | ···················· | ···················· | ···················· | ···················· |
| H | 941 | hCoV-19/Belgium/ULG-10334/2020\|EPI_ISL_540501\|2020 | ···················· | ···················· | ···················· | ···················· | ···················· | ···················· |
| H | 942 | hCoV-19/Belgium/ULG-10335/2020\|EPI_ISL_540502\|2020 | ···················· | ···················· | ···················· | ···················· | ···················· | ···················· |
| H | 943 | hCoV-19/Belgium/ULG-10336/2020\|EPI_ISL_540503\|2020 | ···················· | ···················· | ···················· | ···················· | ···················· | ···················· |
| H | 944 | hCoV-19/Belgium/ULG-10337/2020\|EPI_ISL_540504\|2020 | ···················· | ···················· | ···················· | ···················· | ···················· | ···················· |
| H | 945 | hCoV-19/Belgium/ULG-10338/2020\|EPI_ISL_540505\|2020 | ···················· | ···················· | ···················· | ···················· | ···················· | ···················· |
| H | 946 | hCoV-19/Belgium/ULG-10339/2020\|EPI_ISL_540506\|2020 | ···················· | ···················· | ···················· | ···················· | ···················· | ···················· |
| H | 947 | hCoV-19/Belgium/ULG-10340/2020\|EPI_ISL_540507\|2020 | ···················· | ···················· | ···················· | ···················· | ···················· | ···················· |
| H | 948 | hCoV-19/Belgium/ULG-10341/2020\|EPI_ISL_540508\|2020 | ···················· | ···················· | ···················· | ···················· | ···················· | ···················· |
| H | 949 | hCoV-19/Belgium/ULG-10343/2020\|EPI_ISL_540510\|2020 | ···················· | ···················· | ···················· | ···················· | ···················· | ···················· |
| H | 950 | hCoV-19/Belgium/ULG-10344/2020\|EPI_ISL_540511\|2020 | ···················· | ···················· | ···················· | ···················· | ···················· | ···················· |
| H | 951 | hCoV-19/Belgium/ULG-10345/2020\|EPI_ISL_540512\|2020 | ···················· | ···················· | ···················· | ···················· | ···················· | ···················· |
| H | 952 | hCoV-19/Belgium/ULG-10347/2020\|EPI_ISL_540514\|2020 | ···················· | ···················· | ···················· | ···················· | ···················· | ···················· |
| H | 953 | hCoV-19/Belgium/ULG-10348/2020\|EPI_ISL_540515\|2020 | ···················· | ···················· | ···················· | ···················· | ···················· | ···················· |
| H | 954 | hCoV-19/Belgium/ULG-10350/2020\|EPI_ISL_540517\|2020 | ···················· | ···················· | ···················· | ···················· | ···················· | ···················· |
| H | 955 | hCoV-19/Belgium/ULG-10356/2020\|EPI_ISL_540523\|2020 | ···················· | ···················· | ···················· | ···················· | ···················· | ···················· |
| H | 956 | hCoV-19/Belgium/ULG-10358/2020\|EPI_ISL_540525\|2020 | ···················· | ···················· | ···················· | ···················· | ···················· | ···················· |
| H | 957 | hCoV-19/Belgium/ULG-10360/2020\|EPI_ISL_540527\|2020 | ···················· | ···················· | ···················· | ···················· | ···················· | ···················· |
| H | 958 | hCoV-19/Belgium/ULG-10361/2020\|EPI_ISL_540528\|2020 | ···················· | ···················· | ···················· | ···················· | ···················· | ···················· |
| H | 959 | hCoV-19/Belgium/ULG-10398/2020\|EPI_ISL_540565\|2020 | ···················· | ···················· | ···················· | ···················· | ···················· | ···················· |
| H | 960 | hCoV-19/Belgium/ULG-10399/2020\|EPI_ISL_540566\|2020 | ···················· | ···················· | ···················· | ···················· | ···················· | ···················· |
| H | 961 | hCoV-19/Belgium/ULG-10401/2020\|EPI_ISL_540568\|2020 | ···················· | ···················· | ···················· | ···················· | ···················· | ···················· |
| H | 962 | hCoV-19/Belgium/ULG-10402/2020\|EPI_ISL_540569\|2020 | ···················· | ···················· | ···················· | ···················· | ···················· | ···················· |
| H | 963 | hCoV-19/Belgium/ULG-10406/2020\|EPI_ISL_540573\|2020 | ···················· | ···················· | ···················· | ···················· | ···················· | ···················· |
| H | 964 | hCoV-19/Belgium/ULG-10407/2020\|EPI_ISL_540574\|2020 | ···················· | ···················· | ···················· | ···················· | ···················· | ···················· |
| H | 965 | hCoV-19/Belgium/ULG-10411/2020\|EPI_ISL_540578\|2020 | ···················· | ···················· | ···················· | ···················· | ···················· | ···················· |
| H | 966 | hCoV-19/England/LIVE-AA949/2020\|EPI_ISL_540587\|202 | ···················· | ···················· | ···················· | ···················· | ···················· | ···················· |
| H | 968 | hCoV-19/England/LIVE-AA9A3/2020\|EPI_ISL_540590\|202 | ···················· | ···················· | ···················· | ···················· | ····T··············· | ···················· |
| H | 970 | hCoV-19/England/LIVE-AAA28/2020\|EPI_ISL_540592\|202 | ···················· | ···················· | ···················· | ···················· | ·······NN··········· | ···················· |
| H | 972 | hCoV-19/England/QEUH-998FF8/2020\|EPI_ISL_541810\|20 | ···················· | ···················· | ···················· | ···················· | ···················· | ···················· |
| H | 973 | hCoV-19/England/QEUH-998F25/2020\|EPI_ISL_541811\|20 | ···················· | ···················· | ···················· | ···················· | ···················· | ···················· |
| H | 974 | hCoV-19/England/QEUH-998EBF/2020\|EPI_ISL_541812\|20 | ···················· | ···················· | ···················· | ···················· | ···················· | ···················· |
| H | 975 | hCoV-19/England/QEUH-999276/2020\|EPI_ISL_541813\|20 | ···················· | ···················· | ···················· | ···················· | ···················· | ···················· |
| H | 976 | hCoV-19/England/QEUH-9990E5/2020\|EPI_ISL_541814\|20 | ···················· | ···················· | ···················· | ···················· | ···················· | ···················· |
| H | 977 | hCoV-19/England/QEUH-999407/2020\|EPI_ISL_541815\|20 | ···················· | ···················· | ···················· | ···················· | ···················· | ···················· |
| H | 978 | hCoV-19/England/QEUH-998E91/2020\|EPI_ISL_541816\|20 | ···················· | ·A·················· | ···················· | ···················· | ···················· | ···················· |
| H | 979 | hCoV-19/England/QEUH-998FE9/2020\|EPI_ISL_541818\|20 | ···················· | ···················· | ···················· | ···················· | ···················· | ···················· |
| H | 980 | hCoV-19/England/QEUH-998F07/2020\|EPI_ISL_541819\|20 | ···················· | ·A·················· | ···················· | ···················· | ···················· | ···A················ |
| H | 981 | hCoV-19/England/QEUH-9990F4/2020\|EPI_ISL_541820\|20 | ···················· | ···················· | ···················· | ···················· | ···················· | ···················· |
| H | 982 | hCoV-19/England/QEUH-998FDA/2020\|EPI_ISL_541821\|20 | ···················· | ···················· | ···········T········ | ···················· | ···················· | ···················· |
| H | 983 | hCoV-19/England/QEUH-999319/2020\|EPI_ISL_541823\|20 | ···················· | ···················· | ···················· | ···················· | ···················· | ···················· |
| H | 984 | hCoV-19/England/QEUH-9993EC/2020\|EPI_ISL_541824\|20 | ···················· | ···················· | ···················· | ···················· | ···················· | ···················· |
| H | 985 | hCoV-19/England/QEUH-998F9E/2020\|EPI_ISL_541825\|20 | ···················· | ···················· | ···················· | ···················· | ···················· | ···················· |
| H | 986 | hCoV-19/England/QEUH-998EFB/2020\|EPI_ISL_541826\|20 | ···················· | ···················· | ···················· | ···················· | ···················· | ···················· |
| H | 987 | hCoV-19/England/QEUH-998FCB/2020\|EPI_ISL_541827\|20 | ···················· | ···················· | ···················· | ···················· | ···················· | ···················· |
| H | 988 | hCoV-19/England/QEUH-999188/2020\|EPI_ISL_541828\|20 | ···················· | ···················· | ···················· | ···················· | ···················· | ···················· |
| H | 989 | hCoV-19/England/QEUH-999258/2020\|EPI_ISL_541829\|20 | ···················· | ···················· | ···················· | ···················· | ···················· | ···················· |
| H | 990 | hCoV-19/England/QEUH-999373/2020\|EPI_ISL_541830\|20 | ···················· | ···················· | ···················· | ···················· | ···················· | ···················· |
| H | 991 | hCoV-19/England/QEUH-9992EF/2020\|EPI_ISL_541831\|20 | ···················· | ···················· | ···················· | ···················· | ···················· | ···················· |
| H | 992 | hCoV-19/England/QEUH-998F8F/2020\|EPI_ISL_541832\|20 | ···················· | ···················· | ···················· | ···················· | ···················· | ···················· |
| H | 993 | hCoV-19/Wales/QEUH-998F61/2020\|EPI_ISL_541833\|2020 | ···················· | ···················· | ···················· | ···················· | ···················· | ···················· |
| H | 994 | hCoV-19/England/QEUH-9990D6/2020\|EPI_ISL_541834\|20 | ···················· | ···················· | ···················· | ···················· | ···················· | ···················· |
| H | 995 | hCoV-19/England/QEUH-9993CE/2020\|EPI_ISL_541835\|20 | ···················· | ·A·················· | ···················· | ···················· | ···················· | ···················· |
| H | 996 | hCoV-19/England/QEUH-99906D/2020\|EPI_ISL_541836\|20 | ···················· | ···················· | ···················· | ···················· | ···················· | ···················· |
| H | 997 | hCoV-19/England/QEUH-998F52/2020\|EPI_ISL_541837\|20 | ···················· | ···················· | ···················· | ···················· | ···················· | ···················· |
| H | 998 | hCoV-19/Scotland/QEUH-9990B8/2020\|EPI_ISL_541838\|2 | ···················· | ···················· | ···················· | ···················· | ···················· | ···················· |
| H | 999 | hCoV-19/England/QEUH-99913D/2020\|EPI_ISL_541839\|20 | ···················· | ···················· | ···················· | ···················· | ···················· | ···················· |
| H | 1000 | hCoV-19/England/QEUH-9992B2/2020\|EPI_ISL_541840\|20 | ···················· | ·A·················· | ···················· | ···················· | ···················· | ···A················ |
| H | 1001 | hCoV-19/England/QEUH-999100/2020\|EPI_ISL_541841\|20 | ···················· | ···················· | ···················· | ···················· | ···················· | ···················· |
| H | 1002 | hCoV-19/England/QEUH-999294/2020\|EPI_ISL_541842\|20 | ···················· | ···················· | ···················· | ···················· | ···················· | ···················· |
| H | 1003 | hCoV-19/England/QEUH-998EDD/2020\|EPI_ISL_541844\|20 | ···················· | ···················· | ···················· | ···················· | ···················· | ···················· |

| | | | | | | | | |
|---|---|---|---|---|---|---|---|---|
| H | 1004 | hCoV-19/England/QEUH-99907C/2020\|EPI_ISL_541846\|20 | ···················· | ···················· | ····················· | ···················· | ···················· | ···················· |
| H | 1005 | hCoV-19/Lithuania/MR-LUHS-Eilnr101/2020\|EPI_ISL_54 | ···················· | ···················· | ····················· | ···················· | ···················· | ···················· |
| H | 1006 | hCoV-19/Lithuania/MR-LUHS-Eilnr11/2020\|EPI_ISL_541 | ···················· | ···················· | ····················· | ···················· | ···················· | ···················· |
| H | 1007 | hCoV-19/Lithuania/MR-LUHS-Eilnr12/2020\|EPI_ISL_541 | ···················· | ···················· | ····················· | ···················· | ···················· | ···················· |
| H | 1008 | hCoV-19/Lithuania/MR-LUHS-Eilnr14/2020\|EPI_ISL_541 | ···················· | ···················· | ····················· | ···················· | ···················· | ···················· |
| H | 1009 | hCoV-19/Lithuania/MR-LUHS-Eilnr17/2020\|EPI_ISL_541 | ···················· | ···················· | ····················· | ···················· | ···················· | ···················· |
| H | 1010 | hCoV-19/Lithuania/MR-LUHS-Eilnr25/2020\|EPI_ISL_541 | ···················· | ···················· | ····················· | ···················· | ···················· | ···················· |
| H | 1011 | hCoV-19/Lithuania/MR-LUHS-Eilnr27/2020\|EPI_ISL_541 | ···················· | ···················· | ····················· | ···················· | ···················· | ···················· |
| H | 1012 | hCoV-19/Lithuania/MR-LUHS-Eilnr31/2020\|EPI_ISL_541 | ···················· | ···················· | ····················· | ···················· | ···················· | ···················· |
| H | 1013 | hCoV-19/Lithuania/MR-LUHS-Eilnr33/2020\|EPI_ISL_541 | ···················· | ···················· | ····················· | ···················· | ···················· | ···················· |
| H | 1014 | hCoV-19/Lithuania/MR-LUHS-Eilnr37/2020\|EPI_ISL_541 | ···················· | ···················· | ····················· | ···················· | ···················· | ···················· |
| H | 1015 | hCoV-19/Lithuania/MR-LUHS-Eilnr40/2020\|EPI_ISL_541 | ···················· | ···················· | ····················· | ···················· | ···················· | ···················· |
| H | 1016 | hCoV-19/Lithuania/MR-LUHS-Eilnr43/2020\|EPI_ISL_541 | ···················· | ···················· | ····················· | ···················· | ···················· | ···················· |
| H | 1017 | hCoV-19/Lithuania/MR-LUHS-Eilnr45/2020\|EPI_ISL_541 | ···················· | ···················· | ····················· | ···················· | ···················· | ···················· |
| H | 1018 | hCoV-19/Lithuania/MR-LUHS-Eilnr46/2020\|EPI_ISL_541 | ···················· | ···················· | ····················· | ···················· | ···················· | ···················· |
| H | 1019 | hCoV-19/Lithuania/MR-LUHS-Eilnr49/2020\|EPI_ISL_541 | ···················· | ···················· | ····················· | ···················· | ···················· | ···················· |
| H | 1025 | hCoV-19/England/QEUH-780766/2020\|EPI_ISL_533200\|20 | ···················· | ···················· | ····················· | ···················· | ···················· | ···················· |
| H | 1026 | hCoV-19/Northern Ireland/QEUH-76C5B1/2020\|EPI_ISL_ | ···················· | ···················· | ····················· | ···················· | ···················· | ···················· |
| H | 1027 | hCoV-19/Northern Ireland/QEUH-79CEAE/2020\|EPI_ISL_ | ···················· | ···················· | ····················· | ···················· | ···················· | ···················· |
| H | 1028 | hCoV-19/England/QEUH-795D4E/2020\|EPI_ISL_533203\|20 | ···················· | ···················· | ····················· | ···················· | ···················· | ···················· |
| H | 1029 | hCoV-19/England/QEUH-7A7338/2020\|EPI_ISL_533205\|20 | ···················· | ···················· | ····················· | ···················· | ···················· | ···················· |
| H | 1030 | hCoV-19/England/QEUH-799A16/2020\|EPI_ISL_533206\|20 | ···················· | ···················· | ····················· | ···················· | ···················· | ···················· |
| H | 1031 | hCoV-19/England/QEUH-79D502/2020\|EPI_ISL_533208\|20 | ···················· | ···················· | ····················· | ···················· | ···················· | ···············A···· |
| H | 1032 | hCoV-19/England/QEUH-78C24B/2020\|EPI_ISL_533209\|20 | ···················· | ···················· | ····················· | ···················· | ···················· | ···················· |
| H | 1033 | hCoV-19/England/QEUH-792585/2020\|EPI_ISL_533210\|20 | ···················· | ···················· | ····················· | ···················· | ···················· | ···················· |
| H | 1034 | hCoV-19/England/QEUH-786C70/2020\|EPI_ISL_533211\|20 | ···················· | ···················· | ····················· | ···················· | ···················· | ···················· |
| H | 1035 | hCoV-19/England/QEUH-79D6C3/2020\|EPI_ISL_533212\|20 | ···················· | ···················· | ····················· | ···················· | ···················· | ···················· |
| H | 1036 | hCoV-19/England/QEUH-7927BC/2020\|EPI_ISL_533213\|20 | ···················· | ·A·················· | ····················· | ···················· | ···················· | ···················· |
| H | 1037 | hCoV-19/England/QEUH-77FB5F/2020\|EPI_ISL_533214\|20 | ···················· | ···················· | ····················· | ···················· | ···················· | ···················· |
| H | 1038 | hCoV-19/England/QEUH-7A532B/2020\|EPI_ISL_533215\|20 | ···················· | ···················· | ····················· | ···················· | ···················· | ···················· |
| H | 1039 | hCoV-19/England/QEUH-79301A/2020\|EPI_ISL_533216\|20 | ···················· | ···················· | ····················· | ···················· | ···················· | ···················· |
| H | 1040 | hCoV-19/England/QEUH-7974DE/2020\|EPI_ISL_533217\|20 | ···················· | ···················· | ····················· | ···················· | ···················· | ···················· |
| H | 1041 | hCoV-19/England/QEUH-78FEEA/2020\|EPI_ISL_533218\|20 | ···················· | ···················· | ····················· | ···················· | ···················· | ···················· |
| H | 1042 | hCoV-19/England/QEUH-78D945/2020\|EPI_ISL_533219\|20 | ···················· | ···················· | ····················· | ···················· | ···················· | ···················· |
| H | 1043 | hCoV-19/England/QEUH-7840FE/2020\|EPI_ISL_533220\|20 | ···················· | ···················· | ····················· | ···················· | ···················· | ···················· |
| H | 1044 | hCoV-19/England/QEUH-77B722/2020\|EPI_ISL_533221\|20 | ···················· | ···················· | ····················· | ···················· | ···················· | ···················· |
| H | 1045 | hCoV-19/England/QEUH-5EDFDA/2020\|EPI_ISL_533238\|20 | ···················· | ···················· | ····················· | ···················· | ···················· | ···················· |
| H | 1046 | hCoV-19/England/QEUH-5F0AC4/2020\|EPI_ISL_533239\|20 | ···················· | ···················· | ····················· | ···················· | ···················· | ···················· |
| H | 1047 | hCoV-19/England/QEUH-5F5625/2020\|EPI_ISL_533240\|20 | ···················· | ···················· | ····················· | ···················· | ···················· | ···················· |
| H | 1048 | hCoV-19/Scotland/QEUH-5FBFF7/2020\|EPI_ISL_533241\|2 | ···················· | ···················· | ····················· | ···················· | ···················· | ···················· |
| H | 1049 | hCoV-19/England/QEUH-5EF59A/2020\|EPI_ISL_533243\|20 | ···················· | ···················· | ····················· | ···················· | ···················· | ···················· |
| H | 1050 | hCoV-19/Northern Ireland/QEUH-5EE926/2020\|EPI_ISL_ | ···················· | ···················· | ····················· | ···················· | ···················· | ···················· |
| H | 1051 | hCoV-19/England/QEUH-5FFCCF/2020\|EPI_ISL_533245\|20 | ···················· | ···················· | ····················· | ···················· | ···················· | ···················· |
| H | 1052 | hCoV-19/England/QEUH-5ED3ED/2020\|EPI_ISL_533246\|20 | ···················· | ···················· | ····················· | ···················· | ···················· | ···················· |
| H | 1053 | hCoV-19/England/QEUH-5FD0C5/2020\|EPI_ISL_533247\|20 | ···················· | ···················· | ····················· | ···················· | ···················· | ···················· |
| H | 1054 | hCoV-19/England/QEUH-5F9885/2020\|EPI_ISL_533248\|20 | ···················· | ···················· | ····················· | ···················· | ···················· | ···················· |
| H | 1055 | hCoV-19/England/QEUH-5EF47F/2020\|EPI_ISL_533249\|20 | ···················· | ···················· | ····················· | ···················· | ···················· | ···················· |
| H | 1056 | hCoV-19/Scotland/QEUH-5F8CB7/2020\|EPI_ISL_533252\|2 | ···················· | ···················· | ····················· | ···················· | ···················· | ···················· |
| H | 1057 | hCoV-19/England/QEUH-5F1920/2020\|EPI_ISL_533253\|20 | ···················· | ···················· | ····················· | ···················· | ···················· | ···················· |
| H | 1058 | hCoV-19/England/QEUH-5ED7A5/2020\|EPI_ISL_533254\|20 | ···················· | ···················· | ····················· | ···················· | ···················· | ···················· |
| H | 1059 | hCoV-19/England/QEUH-5EE434/2020\|EPI_ISL_533255\|20 | ···················· | ···················· | ····················· | ···················· | ···················· | ···················· |
| H | 1060 | hCoV-19/Scotland/QEUH-5FBE36/2020\|EPI_ISL_533256\|2 | ···················· | ···················· | ····················· | ···················· | ···················· | ···················· |
| H | 1061 | hCoV-19/England/QEUH-5F0420/2020\|EPI_ISL_533257\|20 | ···················· | ···················· | ····················· | ···················· | ···················· | ···················· |
| H | 1062 | hCoV-19/England/QEUH-5EDAAC/2020\|EPI_ISL_533258\|20 | ···················· | ···················· | ····················· | ···················· | ···················· | ···················· |
| H | 1063 | hCoV-19/England/QEUH-5F1665/2020\|EPI_ISL_533259\|20 | ···················· | ···················· | ····················· | ···················· | ···················· | ···················· |
| H | 1064 | hCoV-19/England/QEUH-5F5616/2020\|EPI_ISL_533260\|20 | ···················· | ···················· | ····················· | ···················· | ···················· | ···················· |
| H | 1065 | hCoV-19/England/QEUH-5F0F01/2020\|EPI_ISL_533261\|20 | ···················· | ···················· | ····················· | ···················· | ···················· | ···················· |
| H | 1066 | hCoV-19/Scotland/QEUH-5F41DA/2020\|EPI_ISL_533262\|2 | ···················· | ···················· | ····················· | ···················· | ···················· | ···················· |
| H | 1067 | hCoV-19/England/QEUH-5FE64A/2020\|EPI_ISL_533263\|20 | ···················· | ···················· | ····················· | ···················· | ···················· | ···················· |
| H | 1068 | hCoV-19/England/QEUH-5EF9E9/2020\|EPI_ISL_533264\|20 | ···················· | ···················· | ····················· | ···················· | ···················· | ···················· |
| H | 1069 | hCoV-19/England/QEUH-5F4E4C/2020\|EPI_ISL_533265\|20 | ···················· | ···················· | ····················· | ···················· | ···················· | ···················· |
| H | 1070 | hCoV-19/Northern Ireland/QEUH-5FF88F/2020\|EPI_ISL_ | ···················· | ···················· | ····················· | ···················· | ···················· | ···················· |
| H | 1071 | hCoV-19/England/QEUH-5FE000/2020\|EPI_ISL_533268\|20 | ···················· | ···················· | ····················· | ···················· | ···················· | ···················· |
| H | 1072 | hCoV-19/England/QEUH-5EF5D6/2020\|EPI_ISL_533269\|20 | ···················· | ···················· | ····················· | ···················· | ···················· | ···················· |
| H | 1073 | hCoV-19/Scotland/QEUH-5F6624/2020\|EPI_ISL_533271\|2 | ···················· | ···················· | ····················· | ···················· | ···················· | ···················· |
| H | 1074 | hCoV-19/England/QEUH-5E819D/2020\|EPI_ISL_533273\|20 | ···················· | ···················· | ····················· | ···················· | ···················· | ···················· |
| H | 1075 | hCoV-19/England/QEUH-5DCC5F/2020\|EPI_ISL_533274\|20 | ···················· | ······A············· | ····················· | ···················· | ···················· | ···················· |
| H | 1076 | hCoV-19/England/QEUH-5DB986/2020\|EPI_ISL_533275\|20 | ···················· | ····A··············· | ····················· | ···················· | ···················· | ···················· |
| H | 1077 | hCoV-19/England/QEUH-5EC357/2020\|EPI_ISL_533277\|20 | ···················· | ···················· | ····················· | ···················· | ···················· | ···················· |
| H | 1078 | hCoV-19/England/QEUH-5DB27C/2020\|EPI_ISL_533278\|20 | ···················· | ···················· | ····················· | ···················· | ···················· | ···················· |
| H | 1079 | hCoV-19/England/QEUH-5E1AB7/2020\|EPI_ISL_533279\|20 | ···················· | ···················· | ····················· | ···················· | ···················· | ···················· |
| H | 1080 | hCoV-19/England/QEUH-5E6AA3/2020\|EPI_ISL_533280\|20 | ···················· | ···················· | ····················· | ···················· | ···················· | ···················· |
| H | 1081 | hCoV-19/Northern Ireland/QEUH-5E535E/2020\|EPI_ISL_ | ···················· | ······A············· | ····················· | ···················· | ···················· | ···················· |
| H | 1082 | hCoV-19/England/QEUH-5E0283/2020\|EPI_ISL_533282\|20 | ···················· | ···················· | ····················· | ···················· | ···················· | ···················· |
| H | 1083 | hCoV-19/Scotland/QEUH-5DD122/2020\|EPI_ISL_533283\|2 | ···················· | ···················· | ····················· | ···················· | ···················· | ···················· |
| H | 1084 | hCoV-19/England/QEUH-5DF032/2020\|EPI_ISL_533284\|20 | ···················· | ···················· | ····················· | ···················· | ···················· | ···················· |
| H | 1085 | hCoV-19/England/QEUH-5DB6DA/2020\|EPI_ISL_533286\|20 | ···················· | ···················· | ····················· | ···················· | ···················· | ···················· |
| H | 1086 | hCoV-19/England/QEUH-5E7F94/2020\|EPI_ISL_533287\|20 | ···················· | ····A··············· | ····················· | ···················· | ···················· | ···················· |
| H | 1087 | hCoV-19/England/QEUH-5DE6B9/2020\|EPI_ISL_533288\|20 | ···················· | ···················· | ····················· | ···················· | ···················· | ···················· |
| H | 1088 | hCoV-19/England/QEUH-5E1307/2020\|EPI_ISL_533290\|20 | ···················· | ···················· | ····················· | ···················· | ···················· | ···················· |

| | | | | | | | | | |
|---|---|---|---|---|---|---|---|---|---|
| H | 1089 | hCoV-19/Wales/QEUH-5E2870/2020\|EPI_ISL_533291\|2020 | ···················· | ···················· | ····················· | ···················· | ···················· | ···················· | |
| H | 1090 | hCoV-19/England/QEUH-5DEBD8/2020\|EPI_ISL_533293\|20 | ···················· | ···················· | ····················· | ···················· | ···················· | ···················· | |
| H | 1091 | hCoV-19/England/QEUH-5E1E06/2020\|EPI_ISL_533294\|20 | ···················· | ···················· | ····················· | ···················· | ···················· | ···················· | |
| H | 1092 | hCoV-19/England/QEUH-5D9FE0/2020\|EPI_ISL_533295\|20 | ···················· | ···················· | ····················· | ···················· | ···················· | ···················· | |
| H | 1093 | hCoV-19/England/QEUH-5EAC7C/2020\|EPI_ISL_533296\|20 | ···················· | ···················· | ····················· | ···················· | ···················· | ···················· | |
| H | 1094 | hCoV-19/England/QEUH-5ECFF9/2020\|EPI_ISL_533297\|20 | ···················· | ···················· | ····················· | ···················· | ···················· | ···················· | |
| H | 1095 | hCoV-19/England/QEUH-5E9A19/2020\|EPI_ISL_533299\|20 | ···················· | ···················A | ····················· | ···················· | ···················· | ···················· | 1 |
| H | 1096 | hCoV-19/Scotland/QEUH-96E508/2020\|EPI_ISL_534500\|2 | ···················· | ···················· | ····················· | ···················· | ···················· | ···················· | |
| H | 1097 | hCoV-19/Scotland/QEUH-96E4CF/2020\|EPI_ISL_534501\|2 | ···················· | ···················· | ····················· | ···················· | ···················· | ···················· | |
| H | 1098 | hCoV-19/Scotland/QEUH-96E1AA/2020\|EPI_ISL_534502\|2 | ···················· | ···················· | ····················· | ···················· | ···················· | ···················· | |
| H | 1099 | hCoV-19/Scotland/QEUH-96E91B/2020\|EPI_ISL_534503\|2 | ···················· | ···················· | ····················· | ···················· | ···················· | ···················· | |
| H | 1100 | hCoV-19/Scotland/QEUH-96E1B9/2020\|EPI_ISL_534505\|2 | ···················· | ···················· | ····················· | ···················· | ···················· | ···················· | |
| H | 1101 | hCoV-19/Scotland/QEUH-96E8E1/2020\|EPI_ISL_534506\|2 | ···················· | ···················· | ····················· | ···················· | ···················· | ···················· | |
| H | 1102 | hCoV-19/Scotland/QEUH-96E1F5/2020\|EPI_ISL_534507\|2 | ···················· | ···················· | ····················· | ···················· | ···················· | ···················· | |
| H | 1103 | hCoV-19/Scotland/QEUH-96E948/2020\|EPI_ISL_534508\|2 | ···················· | ···················· | ····················· | ···················· | ···················· | ···················· | |
| H | 1104 | hCoV-19/Scotland/QEUH-96EB24/2020\|EPI_ISL_534509\|2 | ···················· | ···················· | ····················· | ···················· | ···················· | ···················· | |
| H | 1105 | hCoV-19/Scotland/QEUH-96EB51/2020\|EPI_ISL_534511\|2 | ···················· | ···················· | ····················· | ···················· | ···················· | ···················· | |
| H | 1106 | hCoV-19/Scotland/QEUH-96D93A/2020\|EPI_ISL_534515\|2 | ···················· | ···················· | ····················· | ···················· | ···················· | ···················· | |
| H | 1107 | hCoV-19/Scotland/QEUH-96D448/2020\|EPI_ISL_534516\|2 | ···················· | ···················· | ····················· | ···················· | ···················· | ···················· | |
| H | 1108 | hCoV-19/Scotland/QEUH-96CEC3/2020\|EPI_ISL_534517\|2 | ···················· | ···················· | ····················· | ···················· | ···················· | ···················· | |
| H | 1109 | hCoV-19/Scotland/QEUH-96D0CC/2020\|EPI_ISL_534520\|2 | ···················· | ···················· | ····················· | ···················· | ···················· | ···················· | |
| H | 1110 | hCoV-19/Scotland/QEUH-96D949/2020\|EPI_ISL_534521\|2 | ···················· | ···················· | ····················· | ···················· | ···················· | ···················· | |
| H | 1111 | hCoV-19/Scotland/QEUH-96D967/2020\|EPI_ISL_534522\|2 | ···················· | ···················· | ····················· | ···················· | ···················· | ···················· | |
| H | 1112 | hCoV-19/Scotland/QEUH-96CEF0/2020\|EPI_ISL_534524\|2 | ···················· | ···················· | ····················· | ···················· | ···················· | ···················· | |
| H | 1113 | hCoV-19/Scotland/QEUH-96CF48/2020\|EPI_ISL_534526\|2 | ···················· | ···················· | ····················· | ···················· | ···················· | ···················· | |
| H | 1114 | hCoV-19/Scotland/QEUH-96CF1B/2020\|EPI_ISL_534527\|2 | ···················· | ···················· | ····················· | ···················· | ···················· | ···················· | |
| H | 1115 | hCoV-19/Scotland/QEUH-96CCD8/2020\|EPI_ISL_534528\|2 | ···················· | ···················· | ····················· | ···················· | ···················· | ···················· | |
| H | 1116 | hCoV-19/Scotland/QEUH-96CD7B/2020\|EPI_ISL_534529\|2 | ···················· | ···················· | ····················· | ···················· | ···················· | ···················· | |
| H | 1117 | hCoV-19/Scotland/QEUH-96D78B/2020\|EPI_ISL_534530\|2 | ···················· | ···················· | ····················· | ···················· | ···················· | ···················· | |
| H | 1118 | hCoV-19/Scotland/QEUH-96CCE7/2020\|EPI_ISL_534531\|2 | ···················· | ···················· | ····················· | ···················· | ···················· | ···················· | |
| H | 1119 | hCoV-19/Scotland/QEUH-96D536/2020\|EPI_ISL_534534\|2 | ···················· | ···················· | ····················· | ···················· | ···················· | ···················· | |
| H | 1120 | hCoV-19/Scotland/QEUH-96D484/2020\|EPI_ISL_534536\|2 | ···················· | ···················· | ····················· | ···················· | ···················· | ···················· | |
| H | 1121 | hCoV-19/Scotland/QEUH-96D6E8/2020\|EPI_ISL_534537\|2 | ···················· | ···················· | ····················· | ···················· | ···················· | ···················· | |
| H | 1122 | hCoV-19/Scotland/QEUH-96D91C/2020\|EPI_ISL_534538\|2 | ···················· | ···················· | ····················· | ···················· | ···················· | ···················· | |
| H | 1123 | hCoV-19/Scotland/QEUH-96DD89/2020\|EPI_ISL_534539\|2 | ···················· | ···················· | ····················· | ···················· | ···················· | ···················· | |
| H | 1124 | hCoV-19/Scotland/QEUH-96D642/2020\|EPI_ISL_534541\|2 | ···················· | ···················· | ····················· | ···················· | ···················· | ···················· | |
| H | 1125 | hCoV-19/Scotland/QEUH-96D712/2020\|EPI_ISL_534542\|2 | ···················· | ···················· | ····················· | ···················· | ···················· | ···················· | |
| H | 1126 | hCoV-19/Scotland/QEUH-96DBBC/2020\|EPI_ISL_534544\|2 | ···················· | ···················· | ····················· | ···················· | ···················· | ···················· | |
| H | 1127 | hCoV-19/Scotland/QEUH-96D42A/2020\|EPI_ISL_534545\|2 | ···················· | ···················· | ····················· | ···················· | ···················· | ···················· | |
| H | 1128 | hCoV-19/Scotland/QEUH-96DBAD/2020\|EPI_ISL_534546\|2 | ···················· | ···················· | ····················· | ···················· | ···················· | ···················· | |
| H | 1129 | hCoV-19/Scotland/QEUH-96D5BE/2020\|EPI_ISL_534548\|2 | ···················· | ···················· | ····················· | ···················· | ···················· | ···················· | |
| H | 1130 | hCoV-19/Scotland/QEUH-96D703/2020\|EPI_ISL_534549\|2 | ···················· | ···················· | ····················· | ···················· | ···················· | ···················· | |
| H | 1131 | hCoV-19/Scotland/QEUH-96D475/2020\|EPI_ISL_534551\|2 | ···················· | ···················· | ····················· | ···················· | ···················· | ···················· | |
| H | 1132 | hCoV-19/Scotland/QEUH-96D31E/2020\|EPI_ISL_534552\|2 | ···················· | ···················· | ····················· | ···················· | ···················· | ···················· | |
| H | 1133 | hCoV-19/Scotland/QEUH-96CD6C/2020\|EPI_ISL_534553\|2 | ···················· | ···················· | ····················· | ···················· | ···················· | ···················· | |
| H | 1134 | hCoV-19/Scotland/QEUH-96C87A/2020\|EPI_ISL_534570\|2 | ···················· | ···················· | ····················· | ···················· | ···················· | ···················· | |
| H | 1135 | hCoV-19/Scotland/QEUH-96D4DF/2020\|EPI_ISL_534572\|2 | ···················· | ···················· | ····················· | ···················· | ···················· | ···················· | |
| H | 1136 | hCoV-19/Scotland/QEUH-96CD8A/2020\|EPI_ISL_534573\|2 | ···················· | ···················· | ····················· | ···················· | ···················· | ···················· | |
| H | 1137 | hCoV-19/Scotland/QEUH-96D3D2/2020\|EPI_ISL_534574\|2 | ···················· | ···················· | ····················· | ···················· | ···················· | ···················· | |
| H | 1138 | hCoV-19/Scotland/QEUH-96CFEE/2020\|EPI_ISL_534575\|2 | ···················· | ···················· | ····················· | ···················· | ···················· | ···················· | |
| H | 1139 | hCoV-19/Scotland/QEUH-96CD4E/2020\|EPI_ISL_534576\|2 | ···················· | ···················· | ····················· | ···················· | ···················· | ···················· | |
| H | 1140 | hCoV-19/Scotland/QEUH-96CEB4/2020\|EPI_ISL_534577\|2 | ···················· | ···················· | ····················· | ···················· | ···················· | ···················· | |
| H | 1141 | hCoV-19/Scotland/QEUH-96D800/2020\|EPI_ISL_534578\|2 | ···················· | ···················· | ····················· | ···················· | ···················· | ···················· | |
| H | 1142 | hCoV-19/Scotland/QEUH-96D92B/2020\|EPI_ISL_534579\|2 | ···················· | ···················· | ····················· | ···················· | ···················· | ···················· | |
| H | 1143 | hCoV-19/Scotland/QEUH-96D105/2020\|EPI_ISL_534580\|2 | ···················· | ···················· | ····················· | ···················· | ···················· | ···················· | |
| H | 1144 | hCoV-19/Scotland/QEUH-96D958/2020\|EPI_ISL_534581\|2 | ···················· | ···················· | ····················· | ···················· | ···················· | ···················· | |
| H | 1145 | hCoV-19/Scotland/QEUH-96CDF3/2020\|EPI_ISL_534582\|2 | ···················· | ···················· | ····················· | ···················· | ···················· | ···················· | |
| H | 1146 | hCoV-19/Scotland/QEUH-96D7E5/2020\|EPI_ISL_534583\|2 | ···················· | ···················· | ····················· | ···················· | ···················· | ···················· | |
| H | 1147 | hCoV-19/Scotland/QEUH-96D28A/2020\|EPI_ISL_534584\|2 | ···················· | ···················· | ····················· | ···················· | ···················· | ···················· | |
| H | 1148 | hCoV-19/Scotland/QEUH-96D457/2020\|EPI_ISL_534585\|2 | ···················· | ···················· | ····················· | ···················· | ···················· | ···················· | |
| H | 1149 | hCoV-19/Scotland/QEUH-96CDD5/2020\|EPI_ISL_534586\|2 | ···················· | ···················· | ····················· | ···················· | ···················· | ···················· | |
| H | 1150 | hCoV-19/Scotland/QEUH-96CE0F/2020\|EPI_ISL_534587\|2 | ···················· | ···················· | ····················· | ···················· | ···················· | ···················· | |
| H | 1151 | hCoV-19/Scotland/QEUH-96DDA7/2020\|EPI_ISL_534588\|2 | ···················· | ···················· | ····················· | ···················· | ···················· | ···················· | |
| H | 1152 | hCoV-19/Scotland/QEUH-96CCBA/2020\|EPI_ISL_534589\|2 | ···················· | ···················· | ····················· | ···················· | ···················· | ···················· | |
| H | 1153 | hCoV-19/Scotland/QEUH-96CDA8/2020\|EPI_ISL_534590\|2 | ···················· | ···················· | ····················· | ···················· | ···················· | ···················· | |
| H | 1154 | hCoV-19/Scotland/QEUH-96D879/2020\|EPI_ISL_534591\|2 | ···················· | ···················· | ····················· | ···················· | ···················· | ···················· | |
| H | 1155 | hCoV-19/Scotland/QEUH-96DDB6/2020\|EPI_ISL_534592\|2 | ···················· | ···················· | ····················· | ···················· | ···················· | ···················· | |
| H | 1156 | hCoV-19/Scotland/QEUH-96D40C/2020\|EPI_ISL_534593\|2 | ···················· | ···················· | ····················· | ···················· | ···················· | ···················· | |
| H | 1157 | hCoV-19/Scotland/QEUH-96DCF5/2020\|EPI_ISL_534594\|2 | ···················· | ···················· | ····················· | ···················· | ···················· | ···················· | |
| H | 1158 | hCoV-19/Scotland/QEUH-96D3F0/2020\|EPI_ISL_534595\|2 | ···················· | ···················· | ····················· | ···················· | ···················· | ···················· | |
| H | 1159 | hCoV-19/Scotland/QEUH-96D7F4/2020\|EPI_ISL_534596\|2 | ···················· | ···················· | ····················· | ···················· | ···················· | ···················· | |
| H | 1160 | hCoV-19/Scotland/QEUH-96D721/2020\|EPI_ISL_534597\|2 | ···················· | ···················· | ····················· | ···················· | ···················· | ···················· | |
| H | 1161 | hCoV-19/Scotland/QEUH-96D85B/2020\|EPI_ISL_534598\|2 | ···················· | ···················· | ····················· | ···················· | ···················· | ···················· | |
| H | 1162 | hCoV-19/Scotland/QEUH-96DE1D/2020\|EPI_ISL_534599\|2 | ···················· | ···················· | ····················· | ···················· | ···················· | ···················· | |
| H | 1163 | hCoV-19/Canada/QC-LSPQ-L00221945/2020\|EPI_ISL_5358 | ···················· | ·············A······ | ····················· | ···················· | ···················· | ···················· | |
| H | 1164 | hCoV-19/Canada/QC-LSPQ-L00222020/2020\|EPI_ISL_5358 | ···················· | ···················· | ····················· | ···················· | ···················· | ···················· | |
| H | 1165 | hCoV-19/Canada/QC-LSPQ-L00222035/2020\|EPI_ISL_5358 | ···················· | ···················· | ····················· | ···················· | ···················· | ···················· | |
| H | 1166 | hCoV-19/Canada/QC-LSPQ-L00222448/2020\|EPI_ISL_5358 | ···················· | ···················· | ····················· | ···················· | ···················· | ···················· | |
| H | 1167 | hCoV-19/Canada/QC-LSPQ-L00222575/2020\|EPI_ISL_5358 | ···················· | ···················· | ····················· | ···················· | ···················· | ···················· | |
| H | 1168 | hCoV-19/Canada/QC-LSPQ-L00222731/2020\|EPI_ISL_5358 | ···················· | ···················· | ····················· | ···················· | ···················· | ···················· | |

```
H  1169 hCoV-19/Canada/QC-LSPQ-L00222823/2020|EPI_ISL_5358  ····················  ····················  ····················  ····················  ····················  ····················
H  1170 hCoV-19/Canada/QC-LSPQ-L00224744/2020|EPI_ISL_5358  ····················  ····················  ····················  ····················  ····················  ····················
H  1171 hCoV-19/Canada/QC-LSPQ-L00224745/2020|EPI_ISL_5358  ····················  ····················  ····················  ····················  ····················  ····················
H  1172 hCoV-19/Canada/QC-LSPQ-L00224793/2020|EPI_ISL_5358  ····················  ····················  ····················  ····················  ····················  ····················
H  1173 hCoV-19/Canada/QC-LSPQ-L00224795/2020|EPI_ISL_5358  ····················  ····················  ····················  ····················  ····················  ····················
H  1174 hCoV-19/Canada/QC-LSPQ-L00224812/2020|EPI_ISL_5358  ····················  ····················  ····················  ····················  ····················  ····················
H  1175 hCoV-19/Canada/QC-LSPQ-L00224827/2020|EPI_ISL_5358  ····················  ····················  ····················  ····················  ····················  ····················
H  1176 hCoV-19/Canada/QC-LSPQ-L00224868/2020|EPI_ISL_5358  ····················  ·············A······  ····················  ····················  ····················  ····················
H  1177 hCoV-19/Canada/QC-LSPQ-L00225045/2020|EPI_ISL_5358  ····················  ····················  ····················  ····················  ····················  ····················
H  1178 hCoV-19/Canada/QC-LSPQ-L00225185/2020|EPI_ISL_5358  ····················  ····················  ····················  ····················  ····················  ····················
H  1179 hCoV-19/Canada/QC-LSPQ-L00225484/2020|EPI_ISL_5358  ····················  ····················  ····················  ····················  ····················  ····················
H  1180 hCoV-19/Canada/QC-LSPQ-L00225500/2020|EPI_ISL_5358  ····················  ····················  ····················  ····················  ····················  ····················
H  1181 hCoV-19/Canada/QC-LSPQ-L00225515/2020|EPI_ISL_5358  ····················  ····················  ····················  ····················  ····················  ····················
H  1182 hCoV-19/Canada/QC-LSPQ-L00225655/2020|EPI_ISL_5358  ····················  ····················  ····················  ····················  ····················  ····················
H  1183 hCoV-19/Canada/QC-LSPQ-L00225662/2020|EPI_ISL_5358  ····················  ····················  ····················  ····················  ····················  ····················
H  1184 hCoV-19/Canada/QC-LSPQ-L00225667/2020|EPI_ISL_5358  ····················  ····················  ····················  ····················  ····················  ····················
H  1185 hCoV-19/Canada/QC-LSPQ-L00225685/2020|EPI_ISL_5358  ····················  ····················  ····················  ····················  ····················  ····················
H  1186 hCoV-19/Canada/QC-LSPQ-L00225808/2020|EPI_ISL_5358  ····················  ····················  ····················  ····················  ····················  ····················
H  1187 hCoV-19/Canada/QC-LSPQ-L00225894/2020|EPI_ISL_5358  ····················  ····················  ····················  ····················  ····················  ····················
H  1188 hCoV-19/Canada/QC-LSPQ-L00225956/2020|EPI_ISL_5358  ····················  ····················  ····················  ····················  ····················  ····················
H  1189 hCoV-19/Canada/QC-LSPQ-L00226142/2020|EPI_ISL_5358  ····················  ····················  ····················  ····················  ····················  ····················
H  1190 hCoV-19/Canada/QC-LSPQ-L00226167/2020|EPI_ISL_5358  ····················  ····················  ····················  ····················  ····················  ····················
H  1191 hCoV-19/Canada/QC-LSPQ-L00226624/2020|EPI_ISL_5358  ····················  ····················  ····················  ····················  ····················  ····················
H  1192 hCoV-19/Canada/QC-LSPQ-L00226858/2020|EPI_ISL_5358  ····················  ·A··················  ····················  ····················  ····················  ····················
H  1193 hCoV-19/Canada/QC-LSPQ-L00226938/2020|EPI_ISL_5358  ····················  ····················  ····················  ····················  ····················  ····················
H  1194 hCoV-19/Canada/QC-LSPQ-L00227094/2020|EPI_ISL_5358  ····················  ····················  ····················  ····················  ····················  ····················
H  1195 hCoV-19/Canada/QC-LSPQ-L00227147/2020|EPI_ISL_5358  ····················  ····················  ····················  ····················  ····················  ····················
H  1196 hCoV-19/Canada/QC-LSPQ-L00227476/2020|EPI_ISL_5358  ····················  ····················  ····················  ····················  ····················  ····················
H  1197 hCoV-19/Canada/QC-LSPQ-L00227481/2020|EPI_ISL_5358  ····················  ····················  ····················  ····················  ····················  ····················
H  1198 hCoV-19/Canada/QC-LSPQ-L00227528/2020|EPI_ISL_5358  ····················  ····················  ····················  ····················  ····················  ····················
H  1199 hCoV-19/Canada/QC-LSPQ-L00227658/2020|EPI_ISL_5358  ····················  ····················  ····················  ····················  ····················  ····················
H  1200 hCoV-19/Canada/QC-LSPQ-L00227663/2020|EPI_ISL_5358  ····················  ····················  ····················  ····················  ····················  ····················
H  1201 hCoV-19/Canada/QC-LSPQ-L00227830/2020|EPI_ISL_5358  ····················  ····················  ····················  ····················  ····················  ····················
H  1202 hCoV-19/Canada/QC-LSPQ-L00227996/2020|EPI_ISL_5358  ····················  ····················  ····················  ····················  ····················  ····················
H  1203 hCoV-19/Canada/QC-LSPQ-L00228149/2020|EPI_ISL_5358  ····················  ····················  ····················  ····················  ····················  ····················
H  1204 hCoV-19/Canada/QC-LSPQ-L00228636/2020|EPI_ISL_5358  ····················  ····················  ····················  ····················  ····················  ····················
H  1205 hCoV-19/Canada/QC-LSPQ-L00228687/2020|EPI_ISL_5358  ····················  ····················  ····················  ····················  ····················  ····················
H  1206 hCoV-19/Canada/QC-LSPQ-L00228787/2020|EPI_ISL_5358  ····················  ····················  ····················  ····················  ····················  ····················
H  1207 hCoV-19/Canada/QC-LSPQ-L00228790/2020|EPI_ISL_5358  ····················  ····················  ····················  ····················  ····················  ····················
H  1208 hCoV-19/Canada/QC-LSPQ-L00228862/2020|EPI_ISL_5358  ····················  ····················  ····················  ····················  ····················  ····················
H  1209 hCoV-19/Canada/QC-LSPQ-L00228950/2020|EPI_ISL_5358  ····················  ····················  ····················  ····················  ····················  ····················
H  1210 hCoV-19/Canada/QC-LSPQ-L00228954/2020|EPI_ISL_5358  ····················  ····················  ····················  ····················  ····················  ····················
H  1211 hCoV-19/Canada/QC-LSPQ-L00228964/2020|EPI_ISL_5358  ····················  ····················  ····················  ····················  ····················  ····················
H  1212 hCoV-19/Canada/QC-LSPQ-L00229065/2020|EPI_ISL_5358  ····················  ····················  ····················  ····················  ····················  ····················
H  1213 hCoV-19/Spain/CT-ISCIII-2013593/2020|EPI_ISL_53954  ····················  ····················  ····················  ····················  ····················  ····················
H  1214 hCoV-19/Spain/CT-ISCIII-2013594/2020|EPI_ISL_53954  ····················  ····················  ····················  ····················  ····················  ····················
H  1215 hCoV-19/Spain/CT-ISCIII-2013595/2020|EPI_ISL_53954  ····················  ····················  ····················  ····················  ····················  ····················
H  1216 hCoV-19/Spain/CT-ISCIII-202045/2020|EPI_ISL_539550  ····················  ····················  ····················  ····················  ····················  ····················
H  1217 hCoV-19/Spain/CT-ISCIII-202050/2020|EPI_ISL_539551  ····················  ····················  ····················  ····················  ····················  ····················
H  1218 hCoV-19/Spain/CT-ISCIII-209811/2020|EPI_ISL_539552  ····················  ····················  ····················  ····················  ····················  ····················
H  1219 hCoV-19/Spain/CT-ISCIII-209814/2020|EPI_ISL_539553  ····················  ····················  ····················  ····················  ····················  ····················
H  1220 hCoV-19/Spain/CT-ISCIII-209815/2020|EPI_ISL_539554  ····················  ····················  ····················  ····················  ····················  ····················
H  1221 hCoV-19/Spain/CT-ISCIII-209818/2020|EPI_ISL_539555  ····················  ····················  ····················  ····················  ····················  ····················
H  1222 hCoV-19/Spain/CT-ISCIII-34696/2020|EPI_ISL_539556|  ····················  ····················  ····················  ····················  ····················  ····················
H  1223 hCoV-19/Spain/EX-ISCIII-201239/2020|EPI_ISL_539557  ····················  ····················  ····················  ····················  ····················  ····················
H  1224 hCoV-19/Spain/EX-ISCIII-201248/2020|EPI_ISL_539558  ····················  ····················  ····················  ····················  ····················  ··A·················
H  1225 hCoV-19/Spain/EX-ISCIII-201654/2020|EPI_ISL_539559  ····················  ····················  ····················  ····················  ····················  ····················
H  1226 hCoV-19/Spain/EX-ISCIII-201655/2020|EPI_ISL_539560  ····················  ····················  ····················  ····················  ····················  ····················
H  1227 hCoV-19/Spain/EX-ISCIII-201899/2020|EPI_ISL_539561  ····················  ····················  ····················  ····················  ····················  ····················
H  1228 hCoV-19/Spain/EX-ISCIII-201963/2020|EPI_ISL_539562  ····················  ····················  ····················  ····················  ····················  ····················
H  1229 hCoV-19/Spain/ML-ISCIII-2013686/2020|EPI_ISL_53956  ····················  ····················  ····················  ····················  ····················  ···············A····
H  1230 hCoV-19/Spain/ML-ISCIII-2013688/2020|EPI_ISL_53956  ····················  ····················  ····················  ····················  ····················  ····················
H  1231 hCoV-19/Spain/NA-ISCIII-201944/2020|EPI_ISL_539569  ····················  ····················  ····················  ····················  ····················  ····················
H  1232 hCoV-19/Spain/NA-ISCIII-201945/2020|EPI_ISL_539570  ····················  ····················  ····················  ····················  ····················  ····················
H  1233 hCoV-19/Spain/NA-ISCIII-201951/2020|EPI_ISL_539571  ····················  ····················  ····················  ····················  ····················  ····················
H  1234 hCoV-19/Spain/NA-ISCIII-201952/2020|EPI_ISL_539572  ····················  ····················  ····················  ····················  ····················  ····················
H  1235 hCoV-19/Gabon/ITM-K011/2020|EPI_ISL_539573|2020-03  ····················  ····················  ····················  ····················  ····················  ····················
H  1236 hCoV-19/Gabon/ITM-K023/2020|EPI_ISL_539574|2020-03  ····················  ····················  ····················  ····················  ····················  ····················
H  1237 hCoV-19/Gabon/ITM-K027/2020|EPI_ISL_539575|2020-03  ····················  ····················  ····················  ····················  ····················  ····················
H  1238 hCoV-19/Gabon/ITM-K028/2020|EPI_ISL_539576|2020-03  ····················  ····················  ····················  ····················  ····················  ····················
H  1239 hCoV-19/Germany/NW-HHU-100/2020|EPI_ISL_539577|202  ····················  ····················  ····················  ····················  ····················  ····················
H  1240 hCoV-19/Belgium/ULG-10364/2020|EPI_ISL_540531|2020  ····················  ····················  ····················  ····················  ····················  ····················
H  1241 hCoV-19/Belgium/ULG-10366/2020|EPI_ISL_540533|2020  ····················  ····················  ····················  ····················  ····················  ····················
H  1242 hCoV-19/Belgium/ULG-10367/2020|EPI_ISL_540534|2020  ····················  ····················  ····················  ····················  ····················  ····················
H  1243 hCoV-19/Belgium/ULG-10368/2020|EPI_ISL_540535|2020  ····················  ····················  ····················  ····················  ····················  ····················
H  1244 hCoV-19/Belgium/ULG-10370/2020|EPI_ISL_540537|2020  ····················  ····················  ····················  ····················  ····················  ····················
H  1245 hCoV-19/Belgium/ULG-10374/2020|EPI_ISL_540541|2020  ····················  ····················  ····················  ····················  ····················  ····················
H  1246 hCoV-19/Belgium/ULG-10375/2020|EPI_ISL_540542|2020  ····················  ····················  ····················  ····················  ····················  ····················
H  1247 hCoV-19/Belgium/ULG-10376/2020|EPI_ISL_540543|2020  ····················  ····················  ····················  ····················  ····················  ····················
H  1248 hCoV-19/Belgium/ULG-10377/2020|EPI_ISL_540544|2020  ····················  ····················  ····················  ····················  ····················  ····················
```

```
H 1249 hCoV-19/Belgium/ULG-10380/2020|EPI_ISL_540547|2020 .................... .................... ..................... .................... .................... ....................
H 1250 hCoV-19/Belgium/ULG-10381/2020|EPI_ISL_540548|2020 .................... .................... ..................... .................... .................... ....................
H 1251 hCoV-19/Belgium/ULG-10383/2020|EPI_ISL_540550|2020 .................... .................... ..................... .................... .................... ....................
H 1252 hCoV-19/Belgium/ULG-10384/2020|EPI_ISL_540551|2020 .................... .................... ..................... .................... .................... ....................
H 1253 hCoV-19/Belgium/ULG-10388/2020|EPI_ISL_540555|2020 .................... .................... ..................... .................... .................... ....................
H 1254 hCoV-19/Belgium/ULG-10391/2020|EPI_ISL_540558|2020 .................... .................... ..................... .................... .................... ....................
H 1255 hCoV-19/Belgium/ULG-10392/2020|EPI_ISL_540559|2020 .................... .................... ..................... .................... .................... ....................
H 1256 hCoV-19/Belgium/ULG-10393/2020|EPI_ISL_540560|2020 .................... .................... ..................... .................... .................... ....................
H 1257 hCoV-19/Lithuania/MR-LUHS-Eilnr50/2020|EPI_ISL_541 .................... .................... ..................... .................... .................... ....................
H 1258 hCoV-19/Lithuania/MR-LUHS-Eilnr51/2020|EPI_ISL_541 .................... .................... ..................... .................... .................... ....................
H 1259 hCoV-19/Lithuania/MR-LUHS-Eilnr52/2020|EPI_ISL_541 .................... .................... ..................... .................... .................... ....................
H 1260 hCoV-19/Lithuania/MR-LUHS-Eilnr54/2020|EPI_ISL_541 .................... .................... ..................... .................... .................... ....................
H 1261 hCoV-19/Lithuania/MR-LUHS-Eilnr55/2020|EPI_ISL_541 .................... .................... ..................... .................... .................... ....................
H 1262 hCoV-19/Lithuania/MR-LUHS-Eilnr56/2020|EPI_ISL_541 .................... .................... ..................... .................... .................... ....................
H 1263 hCoV-19/Lithuania/MR-LUHS-Eilnr6/2020|EPI_ISL_5418 .................... .................... ..................... .................... .................... ....................
H 1264 hCoV-19/Lithuania/MR-LUHS-Eilnr65/2020|EPI_ISL_541 .................... .................... ..................... .................... .................... ....................
H 1265 hCoV-19/Lithuania/MR-LUHS-Eilnr7/2020|EPI_ISL_5418 .................... .................... ..................... .................... .................... ....................
H 1266 hCoV-19/Lithuania/MR-LUHS-Eilnr70/2020|EPI_ISL_541 .................... .................... ..................... .................... .................... ....................
H 1267 hCoV-19/Lithuania/MR-LUHS-Eilnr72/2020|EPI_ISL_541 .................... .................... ..................... .................... .................... ....................
H 1268 hCoV-19/Lithuania/MR-LUHS-Eilnr78/2020|EPI_ISL_541 .................... .................... ..................... .................... .................... ....................
H 1269 hCoV-19/Lithuania/MR-LUHS-Eilnr84/2020|EPI_ISL_541 .................... .................... ..................... .................... .................... ....................
H 1270 hCoV-19/Lithuania/MR-LUHS-Eilnr90/2020|EPI_ISL_541 .................... .................... ..................... .................... .................... ....................
H 1271 hCoV-19/Lithuania/MR-LUHS-Eilnr94/2020|EPI_ISL_541 .................... .................... ..................... .................... .................... ....................
H 1272 hCoV-19/Lithuania/MR-LUHS-Eilnr95/2020|EPI_ISL_541 .................... .................... ..................... .................... .................... ....................
H 1274 hCoV-19/Spain/MD-IBV-99007102/2020|EPI_ISL_541884| .................... .................... ..................... .................... .................... ....................
H 1275 hCoV-19/Spain/MD-IBV-99007112/2020|EPI_ISL_541886| .................... .................... ..................... .................... .................... ....................
H 1276 hCoV-19/Spain/MD-IBV-99007113/2020|EPI_ISL_541887| .................... .................... ..................... .................... .................... ....................
H 1281 hCoV-19/England/QEUH-7A9F8D/2020|EPI_ISL_533223|20 .................... .................... ..................... .................... .................... ....................
H 1282 hCoV-19/England/QEUH-794B54/2020|EPI_ISL_533224|20 .................... .................... ..................... .................... .................... ....................
H 1283 hCoV-19/England/QEUH-5F4C15/2020|EPI_ISL_533225|20 .................... .................... ..................... .................... .................... ....................
H 1284 hCoV-19/England/QEUH-5EF1C3/2020|EPI_ISL_533226|20 .................... .................... ..................... .................... .................... ....................
H 1285 hCoV-19/England/QEUH-5F03D8/2020|EPI_ISL_533227|20 .................... .................... ..................... .................... .................... ....................
H 1286 hCoV-19/Scotland/QEUH-5FC442/2020|EPI_ISL_533228|2 .................... .................... ..................... .................... .................... ....................
H 1287 hCoV-19/Scotland/QEUH-5F6466/2020|EPI_ISL_533231|2 .................... .................... ..................... .................... .................... ....................
H 1288 hCoV-19/Scotland/QEUH-5FDA9A/2020|EPI_ISL_533232|2 .................... .................... ..................... .................... .................... ....................
H 1289 hCoV-19/England/QEUH-5F0280/2020|EPI_ISL_533233|20 .................... .................... ..................... .................... .................... ....................
H 1290 hCoV-19/Northern Ireland/QEUH-5FF39D/2020|EPI_ISL_ .................... .................... ..................... .................... .................... ....................
H 1291 hCoV-19/England/QEUH-5F2716/2020|EPI_ISL_533235|20 .................... .................... ..................... .................... .................... ....................
H 1292 hCoV-19/Scotland/QEUH-5FE932/2020|EPI_ISL_533236|2 .................... .................... ..................... .................... .................... ....................
H 1293 hCoV-19/Scotland/QEUH-5FFF8A/2020|EPI_ISL_533237|2 .................... .................... ..................... .................... .................... ....................
H 1294 hCoV-19/Scotland/QEUH-96D493/2020|EPI_ISL_534554|2 .................... .................... ..................... .................... .................... ....................
H 1295 hCoV-19/Scotland/QEUH-96D114/2020|EPI_ISL_534555|2 .................... .................... ..................... .................... .................... ....................
H 1296 hCoV-19/Scotland/QEUH-96D615/2020|EPI_ISL_534556|2 .................... .................... ..................... .................... .................... ....................
H 1297 hCoV-19/Scotland/QEUH-96D74F/2020|EPI_ISL_534557|2 .................... .................... ..................... .................... .................... ....................
H 1298 hCoV-19/Scotland/QEUH-96D34B/2020|EPI_ISL_534558|2 .................... .................... ..................... .................... .................... ....................
H 1299 hCoV-19/Scotland/QEUH-96C810/2020|EPI_ISL_534559|2 .................... .................... ..................... .................... .................... ....................
H 1300 hCoV-19/Scotland/QEUH-96CF75/2020|EPI_ISL_534560|2 .................... .................... ..................... .................... .................... ....................
H 1301 hCoV-19/Scotland/QEUH-96D8E2/2020|EPI_ISL_534562|2 .................... .................... ..................... .................... .................... ....................
H 1302 hCoV-19/Scotland/QEUH-96D68E/2020|EPI_ISL_534563|2 .................... .................... ..................... .................... .................... ....................
H 1303 hCoV-19/Scotland/QEUH-96C8A7/2020|EPI_ISL_534564|2 .................... .................... ..................... .................... .................... ....................
H 1304 hCoV-19/Scotland/QEUH-96D81F/2020|EPI_ISL_534565|2 .................... .................... ..................... .................... .................... ....................
H 1305 hCoV-19/Scotland/QEUH-96D69D/2020|EPI_ISL_534567|2 .................... .................... ..................... .................... .................... ....................
H 1306 hCoV-19/Scotland/QEUH-96D0AE/2020|EPI_ISL_534568|2 .................... .................... ..................... .................... .................... ....................
H 1307 hCoV-19/Scotland/QEUH-96DE2C/2020|EPI_ISL_534601|2 .................... .................... ..................... .................... .................... ....................
H 1308 hCoV-19/Canada/QC-LSPQ-L00229136/2020|EPI_ISL_5358 .................... .................... ..................... .................... .................... ....................
H 1309 hCoV-19/Canada/QC-LSPQ-L00229172/2020|EPI_ISL_5358 .................... .................... ..................... .................... .................... ....................
H 1310 hCoV-19/Canada/QC-LSPQ-L00229212/2020|EPI_ISL_5358 .................... .................... ..................... .................... .................... ....................
H 1311 hCoV-19/Canada/QC-LSPQ-L00229288/2020|EPI_ISL_5358 .................... .................... ..................... .................... .................... ....................
H 1312 hCoV-19/Canada/QC-LSPQ-L00229319/2020|EPI_ISL_5358 .................... .................... ..................... .................... .................... ....................
H 1313 hCoV-19/Canada/QC-LSPQ-L00229326/2020|EPI_ISL_5358 .................... .................... ..................... .................... .................... ....................
H 1314 hCoV-19/Canada/QC-LSPQ-L00229363/2020|EPI_ISL_5358 .................... .................... ..................... .................... .................... ....................
H 1315 hCoV-19/Canada/QC-LSPQ-L00229384/2020|EPI_ISL_5358 .................... .................... ..................... .................... .................... ....................
H 1316 hCoV-19/Canada/QC-LSPQ-L00229410/2020|EPI_ISL_5358 .................... .................... ..................... .................... .................... ....................
H 1317 hCoV-19/Canada/QC-LSPQ-L00229554/2020|EPI_ISL_5358 .................... .................... ..................... .................... .................... ....................
H 1318 hCoV-19/Canada/QC-LSPQ-L00229671/2020|EPI_ISL_5358 .................... .................... ..................... .................... .................... ....................
H 1319 hCoV-19/Canada/QC-LSPQ-L00229727/2020|EPI_ISL_5359 .................... .................... ..................... .................... .................... ....................
H 1320 hCoV-19/Canada/QC-LSPQ-L00229741/2020|EPI_ISL_5359 .................... .................... ..................... .................... .................... ....................
H 1321 hCoV-19/Canada/QC-LSPQ-L00229794/2020|EPI_ISL_5359 .................... .................... ..................... .................... .................... ....................
H 1322 hCoV-19/Canada/QC-LSPQ-L00229796/2020|EPI_ISL_5359 .................... .................... ..................... .................... .................... ....................
H 1323 hCoV-19/Canada/QC-LSPQ-L00229929/2020|EPI_ISL_5359 .................... .................... ..................... .................... .................... ....................
H 1324 hCoV-19/Canada/QC-LSPQ-L00229952/2020|EPI_ISL_5359 .................... .................... ..................... .................... .................... ....................
H 1325 hCoV-19/Canada/QC-LSPQ-L00230020/2020|EPI_ISL_5359 .................... .................... ..................... .................... .................... ....................
H 1326 hCoV-19/Canada/QC-LSPQ-L00230110/2020|EPI_ISL_5359 .................... .................... ..................... .................... .................... ....................
H 1327 hCoV-19/Canada/QC-LSPQ-L00230159/2020|EPI_ISL_5359 .................... .................... ..................... .................... .................... ....................
H 1328 hCoV-19/Canada/QC-LSPQ-L00230189/2020|EPI_ISL_5359 .................... .................... ..................... .................... .................... ....................
H 1329 hCoV-19/Canada/QC-LSPQ-L00230299/2020|EPI_ISL_5359 .................... .................... ..................... .................... .................... ....................
H 1330 hCoV-19/Canada/QC-LSPQ-L00230315/2020|EPI_ISL_5359 .................... .................... ..................... .................... .................... ....................
H 1331 hCoV-19/Canada/QC-LSPQ-L00230319/2020|EPI_ISL_5359 .................... .................... ..................... .................... .................... ....................
H 1332 hCoV-19/Canada/QC-LSPQ-L00230351/2020|EPI_ISL_5359 .................... .................... ..................... .................... .................... ....................
H 1333 hCoV-19/Canada/QC-LSPQ-L00230444/2020|EPI_ISL_5359 .................... .................... ..................... .................... .................... ....................
```

| | | | | | | | | |
|---|---|---|---|---|---|---|---|---|
| H | 1334 | hCoV-19/Canada/QC-LSPQ-L00230448/2020\|EPI_ISL_5359 | ···················· | ···················· | ···················· | ···················· | ···················· | ···················· |
| H | 1335 | hCoV-19/Canada/QC-LSPQ-L00230472/2020\|EPI_ISL_5359 | ···················· | ···················· | ···················· | ···················· | ···················· | ···················· |
| H | 1336 | hCoV-19/Canada/QC-LSPQ-L00230473/2020\|EPI_ISL_5359 | ···················· | ···················· | ···················· | ···················· | ···················· | ···················· |
| H | 1337 | hCoV-19/Canada/QC-LSPQ-L00230482/2020\|EPI_ISL_5359 | ···················· | ···················· | ···················· | ···················· | ···················· | ···················· |
| H | 1338 | hCoV-19/Canada/QC-LSPQ-L00230506/2020\|EPI_ISL_5359 | ···················· | ···················· | ···················· | ···················· | ···················· | ···················· |
| H | 1339 | hCoV-19/Canada/QC-LSPQ-L00230536/2020\|EPI_ISL_5359 | ···················· | ···················· | ···················· | ···················· | ···················· | ···················· |
| H | 1340 | hCoV-19/Canada/QC-LSPQ-L00230637/2020\|EPI_ISL_5359 | ···················· | ···················· | ···················· | ···················· | ···················· | ···················· |
| H | 1341 | hCoV-19/Canada/QC-LSPQ-L00230647/2020\|EPI_ISL_5359 | ···················· | ···················· | ···················· | ···················· | ···················· | ···················· |
| H | 1342 | hCoV-19/Canada/QC-LSPQ-L00230666/2020\|EPI_ISL_5359 | ···················· | ···················· | ···················· | ···················· | ···················· | ···················· |
| H | 1343 | hCoV-19/Canada/QC-LSPQ-L00230676/2020\|EPI_ISL_5359 | ···················· | ···················· | ···················· | ···················· | ···················· | ···················· |
| H | 1344 | hCoV-19/Canada/QC-LSPQ-L00230720/2020\|EPI_ISL_5359 | ···················· | ···················· | ···················· | ···················· | ···················· | ···················· |
| H | 1345 | hCoV-19/Canada/QC-LSPQ-L00230755/2020\|EPI_ISL_5359 | ···················· | ···················· | ···················· | ···················· | ···················· | ···················· |
| H | 1346 | hCoV-19/Canada/QC-LSPQ-L00230787/2020\|EPI_ISL_5359 | ···················· | ···················· | ···················· | ···················· | ···················· | ···················· |
| H | 1347 | hCoV-19/Canada/QC-LSPQ-L00230796/2020\|EPI_ISL_5359 | ···················· | ···················· | ···················· | ···················· | ···················· | ···················· |
| H | 1351 | hCoV-19/Spain/AN-IBV-98001722/2020\|EPI_ISL_538137\| | A··················· | ···················· | ···················· | ···················· | ···················· | ···················· |
| H | 1352 | hCoV-19/Spain/AN-IBV-98001736/2020\|EPI_ISL_538141\| | ···················· | ···················· | ···················· | ···················· | ···················· | ···················· |
| H | 1355 | hCoV-19/Spain/AN-IBV-98001776/2020\|EPI_ISL_538151\| | ···················· | ···················· | ···················· | ···················· | ···················· | ···················· |
| H | 1356 | hCoV-19/Spain/AN-IBV-98001778/2020\|EPI_ISL_538152\| | ···················· | ···················· | ···················· | ···················· | ···················· | ···················· |
| H | 1359 | hCoV-19/Spain/AN-IBV-98001786/2020\|EPI_ISL_538156\| | ···················· | ···················· | ···················· | ···················· | ···················· | ···················· |
| H | 1363 | hCoV-19/Spain/AN-IBV-98001806/2020\|EPI_ISL_538162\| | ···················· | ···················· | ···················· | ···················· | ···················· | ···················· |
| H | 1367 | hCoV-19/Switzerland/VD-ETHZ-260078/2020\|EPI_ISL_53 | ···················· | ···················· | ···················· | ···················· | ···················· | ···················· |
| H | 1368 | hCoV-19/Switzerland/BL-ETHZ-260079/2020\|EPI_ISL_53 | ···················· | ···G················ | ···················· | ···················· | ···················· | ···················· |
| H | 1369 | hCoV-19/Switzerland/AG-ETHZ-260080/2020\|EPI_ISL_53 | ···················· | ···················· | ···················· | ···················· | ···················· | ···················· |
| H | 1370 | hCoV-19/Switzerland/BE-ETHZ-260081/2020\|EPI_ISL_53 | ···················· | ···················· | ···················· | ···················· | ···················· | ···················· |
| H | 1371 | hCoV-19/Switzerland/BE-ETHZ-260084/2020\|EPI_ISL_53 | ···················· | ···················· | ···················· | ···················· | ···················· | ···················· |
| H | 1372 | hCoV-19/Switzerland/BE-ETHZ-260085/2020\|EPI_ISL_53 | ···················· | ···················· | ···················· | ···················· | ···················· | ···················· |
| H | 1373 | hCoV-19/Switzerland/ZH-ETHZ-260089/2020\|EPI_ISL_53 | ···················· | ···················· | ···················· | ···················· | ···················· | ···················· |
| H | 1374 | hCoV-19/Switzerland/VD-ETHZ-260090/2020\|EPI_ISL_53 | ···················· | ···················· | ···················· | ···················· | ···················· | ···················· |
| H | 1375 | hCoV-19/Switzerland/AG-ETHZ-260091/2020\|EPI_ISL_53 | ···················· | ·A·················· | ···················· | ···················· | ···················· | ···················· |
| H | 1376 | hCoV-19/Switzerland/UR-ETHZ-260095/2020\|EPI_ISL_53 | ···········K········ | ·R·················· | ···················· | ···················· | ···················· | ···················· |
| H | 1377 | hCoV-19/Switzerland/VD-ETHZ-260097/2020\|EPI_ISL_53 | ···················· | ···················· | ···················· | ···················· | ···················· | ···················· |
| H | 1378 | hCoV-19/Switzerland/VD-ETHZ-260098/2020\|EPI_ISL_53 | ···················· | ···················· | ···················· | ···················· | ···················· | ···················· |
| H | 1379 | hCoV-19/Switzerland/VD-ETHZ-260099/2020\|EPI_ISL_53 | ···················· | ···················· | ···················· | ···················· | ···················· | ···················· |
| H | 1380 | hCoV-19/Switzerland/BL-ETHZ-260112/2020\|EPI_ISL_53 | ···················· | ···················· | ···················· | ···················· | ······K············· | ···················· |
| H | 1381 | hCoV-19/Switzerland/BL-ETHZ-260113/2020\|EPI_ISL_53 | ···················· | ···················· | ···················· | ···················· | ······K············· | ···················· |
| H | 1382 | hCoV-19/Switzerland/BL-ETHZ-260114/2020\|EPI_ISL_53 | ···················· | ···················· | ···················· | ···················· | ······K············· | ···················· |
| H | 1383 | hCoV-19/Switzerland/BL-ETHZ-260116/2020\|EPI_ISL_53 | ···················· | ···G················ | ···················· | ···················· | ···················· | ···················· |
| H | 1384 | hCoV-19/Switzerland/AG-ETHZ-260117/2020\|EPI_ISL_53 | ···················· | ···················· | ···················· | ···················· | ···················· | ···················· |
| H | 1385 | hCoV-19/Switzerland/ZH-ETHZ-260122/2020\|EPI_ISL_53 | ···················· | ···················· | ···················· | ···················· | ···················· | ···················· |
| H | 1386 | hCoV-19/Switzerland/BE-ETHZ-260169/2020\|EPI_ISL_53 | ···················· | ···················· | ···················· | ···················· | ···················· | ···················· |
| H | 1387 | hCoV-19/Switzerland/BE-ETHZ-260170/2020\|EPI_ISL_53 | ···················· | ···················· | ···················· | ···················· | ···················· | ···················· |
| H | 1388 | hCoV-19/Switzerland/BL-ETHZ-260171/2020\|EPI_ISL_53 | ···················· | ···G················ | ···················· | ···················· | ···················· | ···················· |
| H | 1389 | hCoV-19/Switzerland/SG-ETHZ-260173/2020\|EPI_ISL_53 | ···················· | ···················· | ···················· | ···················· | ···················· | ···················· |
| H | 1390 | hCoV-19/Switzerland/SZ-ETHZ-260174/2020\|EPI_ISL_53 | ···················· | ···················· | ···················· | ···················· | ···················· | ···················· |
| H | 1391 | hCoV-19/Switzerland/VD-ETHZ-260176/2020\|EPI_ISL_53 | ···················· | ···················· | ···················· | ···················· | ···················· | ···················· |
| H | 1392 | hCoV-19/Switzerland/VD-ETHZ-260177/2020\|EPI_ISL_53 | ···················· | ···················· | ···················· | ···················· | ···················· | ···················· |
| H | 1393 | hCoV-19/Switzerland/BE-ETHZ-260180/2020\|EPI_ISL_53 | ···················· | ···················· | ···················· | ···················· | ···················· | ···················· |
| H | 1394 | hCoV-19/Switzerland/VD-ETHZ-260182/2020\|EPI_ISL_53 | ···················· | ···················· | ···················· | ···················· | ···················· | ···················· |
| H | 1395 | hCoV-19/Switzerland/ZH-ETHZ-260184/2020\|EPI_ISL_53 | ···················· | ···················· | ···················· | ···················· | ···················· | ···················· |
| H | 1396 | hCoV-19/Switzerland/ZH-ETHZ-260185/2020\|EPI_ISL_53 | ···················· | ···················· | ···················· | ···················· | ···················· | ···················· |
| H | 1397 | hCoV-19/Switzerland/BE-ETHZ-260187/2020\|EPI_ISL_53 | ···················· | ···················· | ···················· | ···················· | ···················· | ···················· |
| H | 1398 | hCoV-19/Switzerland/SO-ETHZ-260188/2020\|EPI_ISL_53 | ···················· | ···················· | ···················· | ···················· | ······K············· | ···················· |
| H | 1399 | hCoV-19/Switzerland/SO-ETHZ-260190/2020\|EPI_ISL_53 | ···················· | ···················· | ···················· | ···················· | ······K············· | ···················· |
| H | 1400 | hCoV-19/Switzerland/VD-ETHZ-260192/2020\|EPI_ISL_53 | ···················· | ···················· | ···················· | ···················· | ···················· | ···················· |
| H | 1401 | hCoV-19/Switzerland/VD-ETHZ-260195/2020\|EPI_ISL_53 | ···················· | ···················· | ···················· | ···················· | ···················· | ···················· |
| H | 1402 | hCoV-19/Switzerland/VD-ETHZ-260196/2020\|EPI_ISL_53 | ···················· | ···················· | ···················· | ···················· | ···················· | ···················· |
| H | 1403 | hCoV-19/India/PB-IMT-I14/2020\|EPI_ISL_539483\|2020- | ···················· | ···················· | ···················· | ···················· | ···················· | ···················· |
| H | 1404 | hCoV-19/India/PB-IMT-L176/2020\|EPI_ISL_539486\|2020 | ···················· | ···················· | ···················· | ···················· | ···················· | ···················· |
| H | 1405 | hCoV-19/India/PB-IMT-L178/2020\|EPI_ISL_539487\|2020 | ···················· | ···················· | ···················· | ···················· | ···················· | ···················· |
| H | 1406 | hCoV-19/India/UT-IMT-AO15/2020\|EPI_ISL_539491\|2020 | ···················· | ·A·················· | ···················· | ···················· | ···················· | ···A················ |
| H | 1407 | hCoV-19/Czech Republic/NRL-5555/2020\|EPI_ISL_53949 | ···················· | ···················· | ···················· | ···················· | ···················· | ···················· |
| H | 1408 | hCoV-19/Czech Republic/NRL-5635/2020\|EPI_ISL_53949 | ···················· | ···················· | ···················· | ···················· | ···················· | ···················· |
| H | 1409 | hCoV-19/USA/PR-CDC-S29/2020\|EPI_ISL_539495\|2020-07 | ···················· | ···················· | ···················· | ···················· | ···················· | ···················· |
| H | 1410 | hCoV-19/Andorra/202552/2020\|EPI_ISL_539496\|2020-03 | ···················· | ···················· | ···················· | ···················· | ···················· | ···················· |
| H | 1411 | hCoV-19/Spain/AN-ISCIII-201273/2020\|EPI_ISL_539497 | ···················· | ···················· | ···················· | ···················· | ···················· | ···················· |
| H | 1412 | hCoV-19/England/QEUH-98F4D7/2020\|EPI_ISL_540401\|20 | ···················· | ···················· | ···················· | ···················· | ···················· | ···················· |
| H | 1413 | hCoV-19/England/QEUH-98F12B/2020\|EPI_ISL_540402\|20 | ···················· | ···················· | ···················· | ···················· | ···················· | ···················· |
| H | 1414 | hCoV-19/England/QEUH-98F343/2020\|EPI_ISL_540403\|20 | ···················· | ···················· | ···················· | ···················· | ···················· | ···················· |
| H | 1415 | hCoV-19/England/QEUH-98EF22/2020\|EPI_ISL_540404\|20 | ···················· | ···················· | ···················· | ···················· | ···················· | ···················· |
| H | 1416 | hCoV-19/England/QEUH-98F783/2020\|EPI_ISL_540405\|20 | ···················· | ·A·················· | ···················· | ···················· | ···················· | ···················· |
| H | 1417 | hCoV-19/Scotland/QEUH-98F3CB/2020\|EPI_ISL_540406\|2 | ···················· | ···················· | ···················· | ···················· | ···················· | ···················· |
| H | 1418 | hCoV-19/England/QEUH-98F5B6/2020\|EPI_ISL_540407\|20 | ···················· | ···················· | ···················· | ···················· | ···················· | ···················· |
| H | 1419 | hCoV-19/England/QEUH-98EEDA/2020\|EPI_ISL_540408\|20 | ···················· | ···················· | ···················· | ···················· | ···················· | ···················· |
| H | 1420 | hCoV-19/England/QEUH-98F13A/2020\|EPI_ISL_540409\|20 | ···················· | ···················· | ···················· | ···················· | ···················· | ···················· |
| H | 1421 | hCoV-19/England/QEUH-98F255/2020\|EPI_ISL_540414\|20 | ···················· | ···················· | ···················· | ···················· | ···················· | ···················· |
| H | 1422 | hCoV-19/England/QEUH-98EF04/2020\|EPI_ISL_540416\|20 | ···················· | ···················· | ···················· | ···················· | ···················· | ···················· |
| H | 1423 | hCoV-19/England/QEUH-96B046/2020\|EPI_ISL_540419\|20 | ···················· | ···················· | ···················· | ···················· | ···················· | ···················· |
| H | 1424 | hCoV-19/Belgium/ULG-10276/2020\|EPI_ISL_540443\|2020 | ···················· | ···················· | ···················· | ···················· | ···················· | ···················· |
| H | 1425 | hCoV-19/Belgium/ULG-10277/2020\|EPI_ISL_540444\|2020 | ···················· | ···················· | ···················· | ···················· | ···················· | ···················· |
| H | 1426 | hCoV-19/Belgium/ULG-10278/2020\|EPI_ISL_540445\|2020 | ···················· | ···················· | ···················· | ···················· | ···················· | ···················· |

| | | | | | | | | | | |
|---|---|---|---|---|---|---|---|---|---|---|
| H | 1427 | hCoV-19/Belgium/ULG-10279/2020\|EPI_ISL_540446\|2020 | ................... | ................... | ................... | ................... | ................... | ................... | | |
| H | 1428 | hCoV-19/Belgium/ULG-10280/2020\|EPI_ISL_540447\|2020 | ................... | ................... | ................... | ................... | ................... | ................... | | |
| H | 1429 | hCoV-19/Belgium/ULG-10281/2020\|EPI_ISL_540448\|2020 | ................... | ................... | ................... | ................... | ................... | ................... | | |
| H | 1430 | hCoV-19/Belgium/ULG-10282/2020\|EPI_ISL_540449\|2020 | ................... | ................... | ................... | ................... | ................... | ................... | | |
| H | 1431 | hCoV-19/Belgium/ULG-10283/2020\|EPI_ISL_540450\|2020 | ................... | ................... | ................... | ................... | ................... | ................... | | |
| H | 1432 | hCoV-19/Belgium/ULG-10285/2020\|EPI_ISL_540452\|2020 | ................... | ................... | ................... | ................... | ................... | ................... | | |
| H | 1433 | hCoV-19/Belgium/ULG-10286/2020\|EPI_ISL_540453\|2020 | ................... | ................... | ................... | ................... | ................... | ................... | | |
| H | 1434 | hCoV-19/Belgium/ULG-10287/2020\|EPI_ISL_540454\|2020 | ................... | ................... | ................... | ................... | ................... | ................... | | |
| H | 1435 | hCoV-19/Belgium/ULG-10288/2020\|EPI_ISL_540455\|2020 | ................... | ................... | ................... | ................... | ................... | ................... | | |
| H | 1436 | hCoV-19/Belgium/ULG-10290/2020\|EPI_ISL_540457\|2020 | ................... | ................... | ................... | ................... | ................... | ................... | | |
| H | 1437 | hCoV-19/Belgium/ULG-10291/2020\|EPI_ISL_540458\|2020 | ................... | ................... | ................... | ................... | ................... | ................... | | |
| H | 1438 | hCoV-19/Belgium/ULG-10292/2020\|EPI_ISL_540459\|2020 | ................... | ................... | ................... | ................... | ................... | ................... | | |
| H | 1439 | hCoV-19/Belgium/ULG-10293/2020\|EPI_ISL_540460\|2020 | ................... | ................... | ................... | ................... | ................... | ................... | | |
| H | 1440 | hCoV-19/Belgium/ULG-10294/2020\|EPI_ISL_540461\|2020 | ................... | ................... | ................... | ................... | ................... | ................... | | |
| H | 1441 | hCoV-19/Belgium/ULG-10295/2020\|EPI_ISL_540462\|2020 | ................... | ................... | ................... | ................... | ................... | ................... | | |
| H | 1442 | hCoV-19/Belgium/ULG-10296/2020\|EPI_ISL_540463\|2020 | ................... | ................... | ................... | ................... | ................... | ................... | | |
| H | 1443 | hCoV-19/Belgium/ULG-10297/2020\|EPI_ISL_540464\|2020 | ................... | ................... | ................... | ................... | ................... | ................... | | |
| H | 1444 | hCoV-19/Belgium/ULG-10298/2020\|EPI_ISL_540465\|2020 | ................... | ................... | ................... | ................... | ................... | ................... | | |
| H | 1445 | hCoV-19/Belgium/ULG-10299/2020\|EPI_ISL_540466\|2020 | ................... | ................... | ................... | ................... | ................... | ................... | | |
| H | 1446 | hCoV-19/Belgium/ULG-10300/2020\|EPI_ISL_540467\|2020 | ................... | ................... | ................... | ................... | ................... | ................... | | |
| H | 1447 | hCoV-19/Belgium/ULG-10302/2020\|EPI_ISL_540469\|2020 | ................... | ................... | ................... | ................... | ................... | ................... | | |
| H | 1448 | hCoV-19/Belgium/ULG-10303/2020\|EPI_ISL_540470\|2020 | ................... | ................... | ................... | ................... | ................... | ................... | | |
| H | 1449 | hCoV-19/Belgium/ULG-10304/2020\|EPI_ISL_540471\|2020 | ................... | ................... | ................... | ................... | ................... | ................... | | |
| H | 1450 | hCoV-19/Belgium/ULG-10305/2020\|EPI_ISL_540472\|2020 | ................... | ................... | ................... | ................... | ................... | ................... | | |
| H | 1451 | hCoV-19/Belgium/ULG-10306/2020\|EPI_ISL_540473\|2020 | ................... | ................... | ................... | ................... | ................... | ................... | | |
| H | 1452 | hCoV-19/Belgium/ULG-10307/2020\|EPI_ISL_540474\|2020 | ................... | ................... | ................... | ................... | ................... | ................... | | |
| H | 1453 | hCoV-19/Belgium/ULG-10308/2020\|EPI_ISL_540475\|2020 | ................... | ................... | ................... | ................... | ................... | ................... | | |
| H | 1454 | hCoV-19/Belgium/ULG-10309/2020\|EPI_ISL_540476\|2020 | ................... | ................... | ................... | ................... | ................... | ................... | | |
| H | 1455 | hCoV-19/Belgium/ULG-10310/2020\|EPI_ISL_540477\|2020 | ................... | ................... | ................... | ................... | ................... | ................... | | |
| H | 1456 | hCoV-19/Belgium/ULG-10311/2020\|EPI_ISL_540478\|2020 | ................... | ................... | ................... | ................... | ................... | ................... | | |
| H | 1457 | hCoV-19/Belgium/ULG-10312/2020\|EPI_ISL_540479\|2020 | ................... | ................... | ................... | ................... | ................... | ................... | | |
| H | 1458 | hCoV-19/Belgium/ULG-10313/2020\|EPI_ISL_540480\|2020 | ................... | ................... | ................... | ................... | ................... | ................... | | |
| H | 1459 | hCoV-19/Belgium/ULG-10314/2020\|EPI_ISL_540481\|2020 | ................... | ................... | ................... | ................... | ................... | ................... | | |
| H | 1460 | hCoV-19/Belgium/ULG-10315/2020\|EPI_ISL_540482\|2020 | ................... | ................... | ................... | ................... | ................... | ................... | | |
| H | 1461 | hCoV-19/Belgium/ULG-10317/2020\|EPI_ISL_540484\|2020 | ................... | ................... | ................... | ................... | ................... | ................... | | |
| H | 1462 | hCoV-19/Belgium/ULG-10319/2020\|EPI_ISL_540486\|2020 | ................... | ................... | ................... | ................... | ................... | ................... | | |
| H | 1463 | hCoV-19/Belgium/ULG-10320/2020\|EPI_ISL_540487\|2020 | ................... | ................... | ................... | ................... | ................... | ................... | | |
| H | 1464 | hCoV-19/Belgium/ULG-10321/2020\|EPI_ISL_540488\|2020 | ................... | ................... | ................... | ................... | ................... | ................... | | |
| H | 1465 | hCoV-19/Belgium/ULG-10322/2020\|EPI_ISL_540489\|2020 | ................... | ................... | ................... | ................... | ................... | ................... | | |
| H | 1466 | hCoV-19/Belgium/ULG-10323/2020\|EPI_ISL_540490\|2020 | ................... | ................... | ................... | ................... | ................... | ................... | | |
| H | 1467 | hCoV-19/Belgium/ULG-10324/2020\|EPI_ISL_540491\|2020 | ................... | ................... | ................... | ................... | ................... | ................... | | |
| H | 1468 | hCoV-19/Belgium/ULG-10325/2020\|EPI_ISL_540492\|2020 | ................... | ................... | ................... | ................... | ................... | ................... | | |
| H | 1469 | hCoV-19/Belgium/ULG-10326/2020\|EPI_ISL_540493\|2020 | ................... | ................... | ................... | ................... | ................... | ................... | | |
| H | 1470 | hCoV-19/Belgium/ULG-10327/2020\|EPI_ISL_540494\|2020 | ................... | ................... | ................... | ................... | ................... | ................... | | |
| H | 1471 | hCoV-19/Belgium/ULG-10328/2020\|EPI_ISL_540495\|2020 | ................... | ................... | ................... | ................... | ................... | ................... | | |
| H | 1472 | hCoV-19/Belgium/ULG-10329/2020\|EPI_ISL_540496\|2020 | ................... | ................... | ................... | ................... | ................... | ................... | | |
| H | 1473 | hCoV-19/Belgium/ULG-10330/2020\|EPI_ISL_540497\|2020 | ................... | ................... | ................... | ................... | ................... | ................... | | |
| H | 1474 | hCoV-19/Belgium/ULG-10331/2020\|EPI_ISL_540498\|2020 | ................... | ................... | ................... | ................... | ................... | ................... | | |
| H | 1475 | hCoV-19/Belgium/ULG-10332/2020\|EPI_ISL_540499\|2020 | ................... | ................... | ................... | ................... | ................... | ................... | | |
| H | 1476 | hCoV-19/India/MH-NIV-13868/2020\|EPI_ISL_541700\|202 | ................... | ................... | ................... | ................... | ................... | ................... | | |
| H | 1477 | hCoV-19/India/MH-NIV-150015/2020\|EPI_ISL_541701\|20 | ................... | ................... | ................... | ................... | ................... | ................... | | |
| H | 1478 | hCoV-19/India/MH-NIV-150596/2020\|EPI_ISL_541702\|20 | ................... | ................... | ................... | ................... | ................... | ................... | | |
| H | 1479 | hCoV-19/India/MH-NIV-153805/2020\|EPI_ISL_541704\|20 | ................... | ................... | ................... | ................... | ................... | ................... | | |
| H | 1480 | hCoV-19/India/MH-NIV-155669/2020\|EPI_ISL_541705\|20 | ................... | ................... | ................... | ................... | ................... | ................... | | |
| H | 1481 | hCoV-19/India/MH-NIV-156143/2020\|EPI_ISL_541706\|20 | ................... | ................... | ................... | ................... | ................... | ................... | | |
| H | 1485 | hCoV-19/India/MH-NIV-164425/2020\|EPI_ISL_541711\|20 | ................... | ................... | ................... | ................... | ................... | ................... | | |
| H | 1486 | hCoV-19/India/MH-NIV-172607/2020\|EPI_ISL_541713\|20 | ................... | ................... | ................... | ................... | ................... | ................... | | |
| H | 1487 | hCoV-19/India/MH-NIV-173797/2020\|EPI_ISL_541714\|20 | ................... | ................... | ................... | ................... | ................... | ................... | | |
| H | 1488 | hCoV-19/India/MH-NIV-174910/2020\|EPI_ISL_541715\|20 | ................... | ................... | ................... | ................... | ................... | ................... | | |
| H | 1489 | hCoV-19/India/MH-NIV-176041/2020\|EPI_ISL_541716\|20 | ................... | ................... | ................... | ................... | ................... | ................... | | |
| H | 1490 | hCoV-19/India/MH-NIV-176352/2020\|EPI_ISL_541717\|20 | ................... | ................... | ................... | ................... | ................... | ................... | | |
| H | 1491 | hCoV-19/India/MH-NIV-183948/2020\|EPI_ISL_541718\|20 | ................... | ................... | ................... | ................... | ................... | ................... | | |
| H | 1492 | hCoV-19/India/MH-NIV-185628/2020\|EPI_ISL_541719\|20 | .............NNN... | ................... | ................... | ................... | ................... | ................... | | 1 |
| H | 1493 | hCoV-19/India/MH-NIV-186959/2020\|EPI_ISL_541720\|20 | ................... | ................... | ................... | ................... | ................... | ................... | | |
| H | 1494 | hCoV-19/India/MH-NIV-187756/2020\|EPI_ISL_541721\|20 | ................... | ................... | ................... | ................... | ................... | ................... | | |
| H | 1495 | hCoV-19/India/MH-NIV-191205/2020\|EPI_ISL_541723\|20 | ................... | ................... | ................... | ................... | ................... | ................... | | |
| H | 1497 | hCoV-19/India/MH-NIV-52010/2020\|EPI_ISL_541725\|202 | ................... | ................... | ................... | ................... | ................... | ................... | | |
| H | 1500 | hCoV-19/India/MH-NIV-58210/2020\|EPI_ISL_541728\|202 | ................... | ................... | ................... | ................... | ................... | ................... | | |
| H | 1501 | hCoV-19/India/MH-NIV-59437/2020\|EPI_ISL_541729\|202 | ................... | ................... | ................... | ..........GNNNT.... | ................... | ................... | | 1 |
| H | 1503 | hCoV-19/India/MH-NIV-60033/2020\|EPI_ISL_541731\|202 | ................... | ................... | ................... | ........C GTAATN G. | ................... | ................... | 1 | 1 |
| H | 1504 | hCoV-19/India/MH-NIV-69925/2020\|EPI_ISL_541733\|202 | ................... | ................... | ................... | ................... | ................... | ................... | | |
| H | 1505 | hCoV-19/India/MH-NIV-71548/2020\|EPI_ISL_541734\|202 | ................... | ................... | ................... | ................... | ................... | ................... | | |
| H | 1506 | hCoV-19/India/MH-NIV-71791/2020\|EPI_ISL_541735\|202 | ................... | ................... | ................... | ................... | ................... | ................... | | |
| H | 1507 | hCoV-19/India/MH-NIV-72638/2020\|EPI_ISL_541736\|202 | ................... | ................... | ................... | ................... | ................... | ................... | | |
| H | 1508 | hCoV-19/India/MH-NIV-73012/2020\|EPI_ISL_541737\|202 | ................... | ................... | ................... | ................... | ................... | ................... | | |
| H | 1509 | hCoV-19/India/MH-NIV-73500/2020\|EPI_ISL_541738\|202 | ................... | ................... | ................... | ................... | ................... | ................... | | |
| H | 1511 | hCoV-19/India/MH-NIV-74916/2020\|EPI_ISL_541740\|202 | ................... | ................... | ................... | ................... | ................... | ................... | | |
| H | 1512 | hCoV-19/India/MH-NIV-78532/2020\|EPI_ISL_541744\|202 | ................... | ................... | ................... | ................... | ................... | ................... | | |
| H | 1514 | hCoV-19/India/MH-NIV-80896/2020\|EPI_ISL_541746\|202 | ................... | ................... | ................... | ................... | ................... | ................... | | |
| H | 1515 | hCoV-19/India/MH-NIV-81270/2020\|EPI_ISL_541747\|202 | ................... | ................... | ................... | ................... | ................... | ................... | | |

| | | | | | | | | |
|---|---|---|---|---|---|---|---|---|
| H | 1517 | hCoV-19/England/LOND-1286DF7/2020\|EPI_ISL_541775\|2 | ···················· | ···················· | ····················· | ···················· | ···················· | ···················· |
| H | 1518 | hCoV-19/England/LOND-1286DCA/2020\|EPI_ISL_541776\|2 | ···················· | ···················· | ····················· | ···················· | ···················· | ···················· |
| H | 1519 | hCoV-19/England/LOND-1286E5E/2020\|EPI_ISL_541777\|2 | ···················· | ···················· | ····················· | ···················· | ···················· | ···················· |
| H | 1520 | hCoV-19/England/LOND-1286EA9/2020\|EPI_ISL_541778\|2 | ···················· | ···················· | ····················· | ···················· | ···················· | ···················· |
| H | 1521 | hCoV-19/England/LOND-1286E4F/2020\|EPI_ISL_541779\|2 | ···················· | ···················· | ····················· | ···················· | ···················· | ···················· |
| H | 1522 | hCoV-19/England/LOND-1286EC7/2020\|EPI_ISL_541780\|2 | ···················· | ···················· | ····················· | ···················· | ···················· | ···················· |
| H | 1523 | hCoV-19/England/LOND-1286E9A/2020\|EPI_ISL_541781\|2 | ···················· | ···················· | ····················· | ···················· | ···················· | ···················· |
| H | 1524 | hCoV-19/England/LOND-1286D7F/2020\|EPI_ISL_541782\|2 | ···················· | ···················· | ····················· | ···················· | ···················· | ···················· |
| H | 1525 | hCoV-19/England/QEUH-9992A3/2020\|EPI_ISL_541784\|20 | ···················· | ···················· | ····················· | ···················· | ···················· | ···················· |
| H | 1526 | hCoV-19/England/QEUH-998F70/2020\|EPI_ISL_541785\|20 | ···················· | ···················· | ····················· | ···················· | ···················· | ···················· |
| H | 1527 | hCoV-19/England/QEUH-999425/2020\|EPI_ISL_541786\|20 | ···················· | ···················· | ····················· | ···················· | ···················· | ···················· |
| H | 1528 | hCoV-19/Wales/QEUH-998E82/2020\|EPI_ISL_541787\|2020 | ···················· | ···················· | ····················· | ···················· | ···················· | ···················· |
| H | 1529 | hCoV-19/England/QEUH-99930A/2020\|EPI_ISL_541788\|20 | ···················· | ···················· | ····················· | ···················· | ···················· | ···················· |
| H | 1530 | hCoV-19/England/QEUH-999003/2020\|EPI_ISL_541789\|20 | ···················· | ·A·················· | ····················· | ···················· | ···················· | ···A················ |
| H | 1531 | hCoV-19/England/QEUH-9993A0/2020\|EPI_ISL_541790\|20 | ···················· | ···················· | ····················· | ···················· | ···················· | ···················· |
| H | 1532 | hCoV-19/England/QEUH-999030/2020\|EPI_ISL_541791\|20 | ···················· | ·A·················· | ····················· | ···················· | ···················· | ···A················ |
| H | 1533 | hCoV-19/England/QEUH-9993BF/2020\|EPI_ISL_541793\|20 | ···················· | ···················· | ····················· | ···················· | ······T············· | ···················· |
| H | 1534 | hCoV-19/England/QEUH-998EA0/2020\|EPI_ISL_541795\|20 | ···················· | ···················· | ····················· | ···················· | ···················· | ···················· |
| H | 1535 | hCoV-19/England/QEUH-998FBC/2020\|EPI_ISL_541796\|20 | ···················· | ···················· | ····················· | ···················· | ···················· | ···················· |
| H | 1536 | hCoV-19/England/QEUH-999012/2020\|EPI_ISL_541797\|20 | ···················· | ···················· | ····················· | ···················· | ···················· | ···················· |
| H | 1537 | hCoV-19/England/QEUH-96C018/2020\|EPI_ISL_530900\|20 | ···················· | ···················· | ····················· | ···················· | ···················· | ···················· |
| H | 1538 | hCoV-19/England/QEUH-96C124/2020\|EPI_ISL_530901\|20 | ···················· | ···················· | ····················· | ···················· | ···················· | ···················· |
| H | 1539 | hCoV-19/Scotland/QEUH-96C6AD/2020\|EPI_ISL_530902\|2 | ···················· | ···················· | ····················· | ···················· | ···················· | ···················· |
| H | 1540 | hCoV-19/England/QEUH-96C33D/2020\|EPI_ISL_530903\|20 | ···················· | ···················· | ····················· | ···················· | ···················· | ···················· |
| H | 1541 | hCoV-19/Northern Ireland/QEUH-96C5A0/2020\|EPI_ISL_ | ···················· | ···················· | ····················· | ···················· | ···················· | ···················· |
| H | 1542 | hCoV-19/Scotland/QEUH-96C29A/2020\|EPI_ISL_530906\|2 | ···················· | ···················· | ····················· | ···················· | ···················· | ···················· |
| H | 1543 | hCoV-19/England/QEUH-96A85E/2020\|EPI_ISL_530908\|20 | ···················· | ·A·················· | ····················· | ···················· | ···················· | ···A················ |
| H | 1544 | hCoV-19/England/QEUH-969F2D/2020\|EPI_ISL_530909\|20 | ···················· | ···················· | ····················· | ···················· | ···················· | ···················· |
| H | 1545 | hCoV-19/Northern Ireland/QEUH-96C5BF/2020\|EPI_ISL_ | ···················· | ···················· | ····················· | ···················· | ···················· | ···················· |
| H | 1546 | hCoV-19/England/QEUH-96C5EC/2020\|EPI_ISL_530911\|20 | ···················· | ···················· | ····················· | ···················· | ···················· | ···················· |
| H | 1547 | hCoV-19/Northern Ireland/QEUH-96C115/2020\|EPI_ISL_ | ···················· | ···················· | ····················· | ···················· | ···················· | ···················· |
| H | 1548 | hCoV-19/England/QEUH-969E6C/2020\|EPI_ISL_530913\|20 | ···················· | ···················· | ····················· | ···················· | ···················· | ···················· |
| H | 1549 | hCoV-19/Northern Ireland/QEUH-96C027/2020\|EPI_ISL_ | ···················· | ···················· | ····················· | ···················· | ···················· | ···················· |
| H | 1550 | hCoV-19/England/QEUH-96A074/2020\|EPI_ISL_530915\|20 | ···················· | ···················· | ····················· | ···················· | ···················· | ···················· |
| H | 1551 | hCoV-19/England/QEUH-969F4B/2020\|EPI_ISL_530916\|20 | ···················· | ···················· | ····················· | ···················· | ···················· | ···················· |
| H | 1552 | hCoV-19/England/QEUH-969CBD/2020\|EPI_ISL_530917\|20 | ···················· | ···················· | ····················· | ···················· | ···················· | ···················· |
| H | 1553 | hCoV-19/England/QEUH-96A8E5/2020\|EPI_ISL_530919\|20 | ···················· | ···················· | ····················· | ···················· | ···················· | ···················· |
| H | 1554 | hCoV-19/England/QEUH-7A1DE5/2020\|EPI_ISL_533118\|20 | ···················· | ···················· | ····················· | ···················· | ···················· | ···················· |
| H | 1555 | hCoV-19/England/QEUH-79AC4C/2020\|EPI_ISL_533120\|20 | ···················· | ···················· | ····················· | ···················· | ···················· | ···················· |
| H | 1556 | hCoV-19/England/QEUH-799454/2020\|EPI_ISL_533121\|20 | ···················· | ···················· | ····················· | ···················· | ···················· | ···················· |
| H | 1557 | hCoV-19/England/QEUH-78B886/2020\|EPI_ISL_533122\|20 | ···················· | ···················· | ····················· | ···················· | ···················· | ···················· |
| H | 1558 | hCoV-19/England/QEUH-7897D6/2020\|EPI_ISL_533123\|20 | ···················· | ···················· | ····················· | ···················· | ···················· | ···················· |
| H | 1559 | hCoV-19/England/QEUH-797D5B/2020\|EPI_ISL_533124\|20 | ···················· | ···················· | ····················· | ···················· | ···················· | ···················· |
| H | 1560 | hCoV-19/England/QEUH-76BC17/2020\|EPI_ISL_533125\|20 | ···················· | ···················· | ····················· | ···················· | ···················· | ···················· |
| H | 1561 | hCoV-19/England/QEUH-787608/2020\|EPI_ISL_533126\|20 | ···················· | ···················· | ····················· | ···················· | ···················· | ···················· |
| H | 1562 | hCoV-19/England/QEUH-789D2F/2020\|EPI_ISL_533127\|20 | ···················· | ···················· | ····················· | ···················· | ···················· | ···················· |
| H | 1563 | hCoV-19/England/QEUH-790639/2020\|EPI_ISL_533128\|20 | ···················· | ···················· | ····················· | ···················· | ···················· | ···················· |
| H | 1564 | hCoV-19/England/QEUH-76ECD8/2020\|EPI_ISL_533129\|20 | ···················· | ···················· | ····················· | ···················· | ···················· | ···················· |
| H | 1565 | hCoV-19/England/QEUH-795F48/2020\|EPI_ISL_533131\|20 | ···················· | ···················· | ····················· | ···················· | ···················· | ···················· |
| H | 1566 | hCoV-19/Wales/QEUH-79CFBA/2020\|EPI_ISL_533132\|2020 | ···················· | ···················· | ····················· | ···················· | ···················· | ···················· |
| H | 1567 | hCoV-19/England/QEUH-79CA13/2020\|EPI_ISL_533134\|20 | ···················· | ···················· | ····················· | ···················· | ···················· | ···················· |
| H | 1568 | hCoV-19/England/QEUH-79971F/2020\|EPI_ISL_533137\|20 | ···················· | ···················· | ····················· | ···················· | ···················· | ···················· |
| H | 1569 | hCoV-19/England/QEUH-78AC4F/2020\|EPI_ISL_533142\|20 | ···················· | ···················· | ····················· | ···················· | ···················· | ···················· |
| H | 1570 | hCoV-19/England/QEUH-77B82F/2020\|EPI_ISL_533143\|20 | ···················· | ···················· | ····················· | ···················· | ···················· | ···················· |
| H | 1571 | hCoV-19/England/QEUH-79EE60/2020\|EPI_ISL_533145\|20 | ···················· | ···················· | ····················· | ···················· | ···················· | ···················· |
| H | 1572 | hCoV-19/England/QEUH-797B15/2020\|EPI_ISL_533146\|20 | ···················· | ···················· | ····················· | ···················· | ···················· | ···················· |
| H | 1573 | hCoV-19/England/QEUH-796509/2020\|EPI_ISL_533148\|20 | ···················· | ···················· | ····················· | ···················· | ···················· | ···················· |
| H | 1574 | hCoV-19/England/QEUH-76DD6D/2020\|EPI_ISL_533149\|20 | ···················· | ···················· | ····················· | ···················· | ···················· | ···················· |
| H | 1575 | hCoV-19/England/QEUH-7A6ACB/2020\|EPI_ISL_533150\|20 | ···················· | ···················· | ····················· | ···················· | ···················· | ···················· |
| H | 1576 | hCoV-19/Northern Ireland/QEUH-7A4ACD/2020\|EPI_ISL_ | ···················· | ···················· | ····················· | ···················· | ···················· | ···················· |
| H | 1577 | hCoV-19/England/QEUH-7A8F15/2020\|EPI_ISL_533152\|20 | ···················· | ···················· | ····················· | ···················· | ···················· | ···················· |
| H | 1578 | hCoV-19/England/QEUH-799B9B/2020\|EPI_ISL_533153\|20 | ···················· | ···················· | ····················· | ···················· | ···················· | ···················· |
| H | 1579 | hCoV-19/England/QEUH-78E777/2020\|EPI_ISL_533155\|20 | ···················· | ···················· | ····················· | ···················· | ···················· | ···················· |
| H | 1580 | hCoV-19/England/QEUH-79BF33/2020\|EPI_ISL_533156\|20 | ···················· | ···················· | ····················· | ···················· | ···················· | ···················· |
| H | 1581 | hCoV-19/England/QEUH-785C71/2020\|EPI_ISL_533157\|20 | ···················· | ···················· | ····················· | ···················· | ···················· | ···················· |
| H | 1582 | hCoV-19/England/QEUH-79AFE9/2020\|EPI_ISL_533158\|20 | ···················· | ···················· | ····················· | ···················· | ···················· | ···················· |
| H | 1583 | hCoV-19/England/QEUH-76D073/2020\|EPI_ISL_533159\|20 | ···················· | ···················· | ····················· | ···················· | ···················· | ···················· |
| H | 1584 | hCoV-19/England/QEUH-76BF2D/2020\|EPI_ISL_533160\|20 | ···················· | ···················· | ····················· | ···················· | ···················· | ···················· |
| H | 1585 | hCoV-19/Northern Ireland/QEUH-79CECC/2020\|EPI_ISL_ | ···················· | ···················· | ····················· | ···················· | ···················· | ···················· |
| H | 1586 | hCoV-19/England/QEUH-797F73/2020\|EPI_ISL_533162\|20 | ···················· | ···················· | ····················· | ···················· | ···················· | ···················· |
| H | 1587 | hCoV-19/England/QEUH-780AF4/2020\|EPI_ISL_533163\|20 | ···················· | ···················· | ····················· | ···················· | ···················· | ···················· |
| H | 1588 | hCoV-19/England/QEUH-795DB7/2020\|EPI_ISL_533164\|20 | ···················· | ···················· | ····················· | ···················· | ···················· | ···················· |
| H | 1589 | hCoV-19/England/QEUH-798EEE/2020\|EPI_ISL_533165\|20 | ···················· | ···················· | ····················· | ···················· | ···················· | ···················· |
| H | 1590 | hCoV-19/England/QEUH-79C5B8/2020\|EPI_ISL_533167\|20 | ···················· | ···················· | ····················· | ···················· | ···················· | ···················· |
| H | 1591 | hCoV-19/England/QEUH-7858C8/2020\|EPI_ISL_533168\|20 | ···················· | ···················· | ····················· | ···················· | ···················· | ···················· |
| H | 1592 | hCoV-19/Northern Ireland/QEUH-77DB9D/2020\|EPI_ISL_ | ···················· | ···················· | ····················· | ···················· | ···················· | ···················· |
| H | 1593 | hCoV-19/England/QEUH-7A84F8/2020\|EPI_ISL_533170\|20 | ···················· | ···················· | ····················· | ···················· | ···················· | ···················· |
| H | 1594 | hCoV-19/England/QEUH-79F00F/2020\|EPI_ISL_533171\|20 | ···················· | ···················· | ····················· | ···················· | ···················· | ···················· |
| H | 1595 | hCoV-19/England/QEUH-77ADF4/2020\|EPI_ISL_533172\|20 | ···················· | ···················· | ····················· | ···················· | ···················· | ···················· |
| H | 1596 | hCoV-19/England/QEUH-79F04B/2020\|EPI_ISL_533173\|20 | ···················· | ···················· | ····················· | ···················· | ···················· | ···················· |

```
H 1597 hCoV-19/England/QEUH-787C60/2020|EPI_ISL_533174|20 ···················· ···················· ····················· ···················· ···················· ····················
H 1598 hCoV-19/England/QEUH-789A28/2020|EPI_ISL_533176|20 ···················· ···················· ····················· ···················· ···················· ····················
H 1599 hCoV-19/Northern Ireland/QEUH-7A4D4C/2020|EPI_ISL_ ···················· ···················· ····················· ···················· ···················· ····················
H 1600 hCoV-19/England/QEUH-78D21D/2020|EPI_ISL_533178|20 ···················· ···················· ····················· ···················· ···················· ····················
H 1601 hCoV-19/England/QEUH-79F807/2020|EPI_ISL_533179|20 ···················· ···················· ····················· ···················· ···················· ····················
H 1602 hCoV-19/England/QEUH-79EE7F/2020|EPI_ISL_533181|20 ···················· ·A·················· ····················· ···················· ···················· ····················
H 1603 hCoV-19/England/QEUH-787301/2020|EPI_ISL_533182|20 ···················· ···················· ····················· ···················· ···················· ····················
H 1604 hCoV-19/England/QEUH-78886B/2020|EPI_ISL_533183|20 ···················· ···················· ····················· ···················· ···················· ····················
H 1605 hCoV-19/England/QEUH-7885A0/2020|EPI_ISL_533184|20 ···················· ···················· ···········T········· ···················· ···················· ····················
H 1606 hCoV-19/England/QEUH-791340/2020|EPI_ISL_533185|20 ···················· ···················· ····················· ···················· ···················· ····················
H 1607 hCoV-19/England/QEUH-79B407/2020|EPI_ISL_533186|20 ···················· ···················· ····················· ···················· ···················· ····················
H 1608 hCoV-19/England/QEUH-79C8EC/2020|EPI_ISL_533187|20 ···················· ···················· ····················· ···················· ···················· ····················
H 1609 hCoV-19/England/QEUH-785567/2020|EPI_ISL_533189|20 ···················· ···················· ····················· ···················· ···················· ····················
H 1610 hCoV-19/Northern Ireland/QEUH-77BCE7/2020|EPI_ISL_ ···················· ···················· ····················· ···················· ···················· ····················
H 1611 hCoV-19/England/QEUH-79F694/2020|EPI_ISL_533192|20 ···················· ···················· ····················· ···················· ···················· ····················
H 1612 hCoV-19/England/QEUH-784638/2020|EPI_ISL_533193|20 ···················· ···················· ····················· ···················· ···················· ····················
H 1613 hCoV-19/England/QEUH-7A7426/2020|EPI_ISL_533194|20 ···················· ···················· ····················· ···················· ···················· ····················
H 1614 hCoV-19/England/QEUH-79EED9/2020|EPI_ISL_533195|20 ···················· ·A·················· ····················· ···················· ···················· ····················
H 1615 hCoV-19/England/QEUH-7948D5/2020|EPI_ISL_533196|20 ···················· ·A·················· ····················· ···················· ···················· ····················
H 1616 hCoV-19/England/QEUH-79FA4D/2020|EPI_ISL_533197|20 ···················· ···················· ····················· ···················· ···················· ····················
H 1617 hCoV-19/England/QEUH-796554/2020|EPI_ISL_533198|20 ···················· ···················· ····················· ···················· ···················· ····················
H 1618 hCoV-19/Scotland/QEUH-9905B2/2020|EPI_ISL_534449|2 ···················· ·A·················· ····················· ···················· ···················· ··A·················
H 1619 hCoV-19/Scotland/QEUH-96E17D/2020|EPI_ISL_534452|2 ···················· ···················· ····················· ···················· ···················· ····················
H 1620 hCoV-19/Scotland/QEUH-96E386/2020|EPI_ISL_534453|2 ···················· ···················· ····················· ···················· ···················· ····················
H 1621 hCoV-19/Scotland/QEUH-96E85A/2020|EPI_ISL_534455|2 ···················· ···················· ····················· ···················· ···················· ····················
H 1622 hCoV-19/Scotland/QEUH-96E8A5/2020|EPI_ISL_534456|2 ···················· ···················· ····················· ···················· ···················· ····················
H 1623 hCoV-19/Scotland/QEUH-96E33B/2020|EPI_ISL_534457|2 ···················· ···················· ····················· ···················· ···················· ····················
H 1624 hCoV-19/Scotland/QEUH-96E447/2020|EPI_ISL_534458|2 ···················· ···················· ····················· ···················· ···················· ····················
H 1625 hCoV-19/Scotland/QEUH-96EA54/2020|EPI_ISL_534459|2 ···················· ···················· ····················· ···················· ···················· ····················
H 1626 hCoV-19/Scotland/QEUH-96E553/2020|EPI_ISL_534461|2 ···················· ···················· ····················· ···················· ···················· ····················
H 1627 hCoV-19/Scotland/QEUH-96EA63/2020|EPI_ISL_534462|2 ···················· ···················· ····················· ···················· ···················· ····················
H 1628 hCoV-19/Scotland/QEUH-96E517/2020|EPI_ISL_534463|2 ···················· ···················· ····················· ···················· ···················· ····················
H 1629 hCoV-19/Scotland/QEUH-96EBE8/2020|EPI_ISL_534465|2 ···················· ···················· ····················· ···················· ···················· ····················
H 1630 hCoV-19/Scotland/QEUH-96E4FC/2020|EPI_ISL_534467|2 ···················· ···················· ····················· ···················· ···················· ····················
H 1631 hCoV-19/Scotland/QEUH-96E8B4/2020|EPI_ISL_534468|2 ···················· ···················· ····················· ···················· ···················· ····················
H 1632 hCoV-19/Scotland/QEUH-96E1C8/2020|EPI_ISL_534469|2 ···················· ···················· ····················· ···················· ···················· ····················
H 1633 hCoV-19/Scotland/QEUH-96E975/2020|EPI_ISL_534470|2 ···················· ···················· ····················· ···················· ···················· ····················
H 1634 hCoV-19/Scotland/QEUH-96E66F/2020|EPI_ISL_534471|2 ···················· ···················· ····················· ···················· ···················· ····················
H 1635 hCoV-19/Scotland/QEUH-96EB15/2020|EPI_ISL_534472|2 ···················· ···················· ····················· ···················· ···················· ····················
H 1636 hCoV-19/Scotland/QEUH-96EB8E/2020|EPI_ISL_534473|2 ···················· ···················· ····················· ···················· ···················· ····················
H 1637 hCoV-19/Scotland/QEUH-96E984/2020|EPI_ISL_534474|2 ···················· ···················· ····················· ···················· ···················· ····················
H 1638 hCoV-19/Scotland/QEUH-96E26B/2020|EPI_ISL_534476|2 ···················· ···················· ····················· ···················· ···················· ····················
H 1639 hCoV-19/Scotland/QEUH-96ECC7/2020|EPI_ISL_534477|2 ···················· ···················· ····················· ···················· ···················· ····················
H 1640 hCoV-19/Scotland/QEUH-96F024/2020|EPI_ISL_534478|2 ···················· ···················· ····················· ···················· ···················· ····················
H 1641 hCoV-19/Scotland/QEUH-96EF28/2020|EPI_ISL_534480|2 ···················· ···················· ····················· ···················· ···················· ····················
H 1642 hCoV-19/Scotland/QEUH-96E32C/2020|EPI_ISL_534481|2 ···················· ···················· ····················· ···················· ···················· ····················
H 1643 hCoV-19/Scotland/QEUH-96E939/2020|EPI_ISL_534482|2 ···················· ···················· ····················· ···················· ···················· ····················
H 1644 hCoV-19/Scotland/QEUH-96E6AB/2020|EPI_ISL_534483|2 ···················· ···················· ····················· ···················· ···················· ····················
H 1645 hCoV-19/Scotland/QEUH-96ECD6/2020|EPI_ISL_534484|2 ···················· ···················· ····················· ···················· ···················· ····················
H 1646 hCoV-19/Scotland/QEUH-96EF64/2020|EPI_ISL_534486|2 ···················· ···················· ····················· ···················· ···················· ····················
H 1647 hCoV-19/Scotland/QEUH-96E526/2020|EPI_ISL_534487|2 ···················· ·A·················· ····················· ···················· ···················· ····················
H 1648 hCoV-19/Scotland/QEUH-96E957/2020|EPI_ISL_534488|2 ···················· ···················· ····················· ···················· ···················· ····················
H 1649 hCoV-19/Scotland/QEUH-96E429/2020|EPI_ISL_534489|2 ···················· ···················· ····················· ···················· ···················· ····················
H 1650 hCoV-19/Scotland/QEUH-96EDE2/2020|EPI_ISL_534490|2 ···················· ···················· ····················· ···················· ···················· ····················
H 1651 hCoV-19/Scotland/QEUH-96EBBB/2020|EPI_ISL_534491|2 ···················· ···················· ····················· ···················· ···················· ····················
H 1652 hCoV-19/Scotland/QEUH-96E544/2020|EPI_ISL_534493|2 ···················· ···················· ····················· ···················· ···················· ····················
H 1653 hCoV-19/Scotland/QEUH-96E298/2020|EPI_ISL_534495|2 ···················· ···················· ····················· ···················· ···················· ····················
H 1654 hCoV-19/Scotland/QEUH-96EF73/2020|EPI_ISL_534496|2 ···················· ···················· ····················· ···················· ···················· ····················
H 1655 hCoV-19/Scotland/QEUH-96E84B/2020|EPI_ISL_534497|2 ···················· ···················· ····················· ···················· ···················· ····················
H 1656 hCoV-19/Scotland/QEUH-96E1D7/2020|EPI_ISL_534498|2 ···················· ···················· ····················· ···················· ···················· ····················
H 1657 hCoV-19/Scotland/QEUH-96E67E/2020|EPI_ISL_534499|2 ···················· ·A·················· ····················· ···················· ···················· ····················
H 1658 hCoV-19/USA/CA-CDC-4729/2020|EPI_ISL_535700|2020-0 ···················· ·A·················· ····················· ···················· ···················· ····················
H 1659 hCoV-19/USA/CA-CDC-4730/2020|EPI_ISL_535701|2020-0 ···················· ·A·················· ····················· ···················· ··C················· ····················
H 1660 hCoV-19/USA/CA-CDC-4731/2020|EPI_ISL_535702|2020-0 ···················· ·A·················· ····················· ···················· ···················· ····················
H 1661 hCoV-19/USA/CA-CDC-4733/2020|EPI_ISL_535703|2020-0 ···················· ·A·················· ····················· ···················· ···················· ····················
H 1662 hCoV-19/USA/CA-CDC-4735/2020|EPI_ISL_535704|2020-0 ···················· ·A·················· ····················· ···················· ···················· ····················
H 1663 hCoV-19/USA/CA-CDC-4739/2020|EPI_ISL_535705|2020-0 ···················· ·A·················· ····················· ···················· ···················· ····················
H 1664 hCoV-19/USA/CA-CDC-4740/2020|EPI_ISL_535706|2020-0 ···················· ·A·················· ····················· ···················· ···················· ····················
H 1665 hCoV-19/USA/CA-CDC-4745/2020|EPI_ISL_535707|2020-0 ···················· ·A·················· ····················· ···················· ···················· ····················
H 1666 hCoV-19/USA/CA-CDC-4746/2020|EPI_ISL_535708|2020-0 ···················· ·A·················· ····················· ···················· ···················· ····················
H 1667 hCoV-19/USA/CA-CDC-4747/2020|EPI_ISL_535709|2020-0 ···················· ·A·················· ····················· ···················· ···················· ····················
H 1668 hCoV-19/USA/CA-CDC-4748/2020|EPI_ISL_535710|2020-0 ···················· ·A·················· ····················· ···················· ···················· ····················
H 1669 hCoV-19/USA/CA-CDC-4749/2020|EPI_ISL_535711|2020-0 ···················· ·A·················· ····················· ···················· ···················· ····················
H 1670 hCoV-19/USA/CA-CDC-4750/2020|EPI_ISL_535712|2020-0 ···················· ·A·················· ····················· ···················· ···················· ····················
H 1671 hCoV-19/USA/CA-CDC-4751/2020|EPI_ISL_535713|2020-0 ···················· ·A·················· ····················· ···················· ···················· ····················
H 1672 hCoV-19/USA/CA-CDC-4752/2020|EPI_ISL_535714|2020-0 ···················· ·A·················· ····················· ···················· ···················· ····················
H 1673 hCoV-19/USA/CA-CDC-4753/2020|EPI_ISL_535715|2020-0 ···················· ·A·················· ····················· ···················· ···················· ····················
H 1674 hCoV-19/Canada/QC-LSPQ-L00210314/2020|EPI_ISL_5357 ···················· ·A·················· ····················· ···················· ···················· ····················
H 1675 hCoV-19/Canada/QC-LSPQ-L00212401/2020|EPI_ISL_5357 ···················· ···················· ····················· ···················· ···················· ····················
H 1676 hCoV-19/Canada/QC-LSPQ-L00213013/2020|EPI_ISL_5357 ···················· ···················· ····················· ···················· ···················· ····················
```

| | | | | | | | | |
|---|---|---|---|---|---|---|---|---|
| H | 1677 | hCoV-19/Canada/QC-LSPQ-L00213312/2020\|EPI_ISL_5357 | ···················· | ···················· | ···················· | ···················· | ···················· | ···················· |
| H | 1678 | hCoV-19/Canada/QC-LSPQ-L00213733/2020\|EPI_ISL_5357 | ···················· | ···················· | ···················· | ···················· | ···················· | ···················· |
| H | 1679 | hCoV-19/Canada/QC-LSPQ-L00213861/2020\|EPI_ISL_5357 | ···················· | ···················· | ···················· | ···················· | ···················· | ···················· |
| H | 1680 | hCoV-19/Canada/QC-LSPQ-L00213981/2020\|EPI_ISL_5357 | ···················· | ···················· | ···················· | ···················· | ···················· | ···················· |
| H | 1681 | hCoV-19/Canada/QC-LSPQ-L00214023/2020\|EPI_ISL_5357 | ···················· | ···················· | ···················· | ···················· | ···················· | ···················· |
| H | 1682 | hCoV-19/Canada/QC-LSPQ-L00214050/2020\|EPI_ISL_5357 | ···················· | ···················· | ···················· | ···················· | ···················· | ···················· |
| H | 1683 | hCoV-19/Canada/QC-LSPQ-L00214081/2020\|EPI_ISL_5357 | ···················· | ···················· | ···················· | ···················· | ···················· | ···················· |
| H | 1684 | hCoV-19/Canada/QC-LSPQ-L00214237/2020\|EPI_ISL_5357 | ···················· | ···················· | ···················· | ···················· | ···················· | ···················· |
| H | 1685 | hCoV-19/Canada/QC-LSPQ-L00214488/2020\|EPI_ISL_5357 | ···················· | ···················· | ···················· | ···················· | ···················· | ···················· |
| H | 1686 | hCoV-19/Canada/QC-LSPQ-L00214517/2020\|EPI_ISL_5357 | ···················· | ···················· | ···················· | ···················· | ···················· | ···················· |
| H | 1687 | hCoV-19/Canada/QC-LSPQ-L00214546/2020\|EPI_ISL_5357 | ···················· | ···················· | ···················· | ···················· | ···················· | ···················· |
| H | 1688 | hCoV-19/Canada/QC-LSPQ-L00214634/2020\|EPI_ISL_5357 | ···················· | ···················· | ···················· | ···················· | ···················· | ···················· |
| H | 1689 | hCoV-19/Canada/QC-LSPQ-L00218353/2020\|EPI_ISL_5357 | ···················· | ···················· | ···················· | ···················· | ···················· | ···················· |
| H | 1690 | hCoV-19/Canada/QC-LSPQ-L00218371/2020\|EPI_ISL_5357 | ···················· | ···················· | ···················· | ···················· | ···················· | ···················· |
| H | 1691 | hCoV-19/Canada/QC-LSPQ-L00218472/2020\|EPI_ISL_5357 | ···················· | ···················· | ···················· | ···················· | ···················· | ···················· |
| H | 1692 | hCoV-19/Canada/QC-LSPQ-L00218475/2020\|EPI_ISL_5357 | ···················· | ···················· | ···················· | ···················· | ···················· | ···················· |
| H | 1693 | hCoV-19/Canada/QC-LSPQ-L00218477/2020\|EPI_ISL_5357 | ···················· | ···················· | ···················· | ···················· | ···················· | ···················· |
| H | 1694 | hCoV-19/Canada/QC-LSPQ-L00218534/2020\|EPI_ISL_5357 | ···················· | ···················· | ···················· | ···················· | ···················· | ···················· |
| H | 1695 | hCoV-19/Canada/QC-LSPQ-L00218559/2020\|EPI_ISL_5357 | ···················· | ···················· | ···················· | ···················· | ···················· | ···················· |
| H | 1696 | hCoV-19/Canada/QC-LSPQ-L00219020/2020\|EPI_ISL_5357 | ···················· | ···················· | ···················· | ···················· | ···················· | ···················· |
| H | 1697 | hCoV-19/Canada/QC-LSPQ-L00219039/2020\|EPI_ISL_5357 | ···················· | ···················· | ···················· | ···················· | ···················· | ···················· |
| H | 1698 | hCoV-19/Canada/QC-LSPQ-L00219049/2020\|EPI_ISL_5357 | ···················· | ···················· | ···················· | ···················· | ···················· | ···················· |
| H | 1699 | hCoV-19/Canada/QC-LSPQ-L00219066/2020\|EPI_ISL_5357 | ···················· | ···················· | ···················· | ···················· | ···················· | ···················· |
| H | 1700 | hCoV-19/Canada/QC-LSPQ-L00219089/2020\|EPI_ISL_5357 | ···················· | ···················· | ···················· | ···················· | ···················· | ···················· |
| H | 1701 | hCoV-19/Canada/QC-LSPQ-L00219335/2020\|EPI_ISL_5357 | ···················· | ···················· | ···················· | ···················· | ···················· | ···················· |
| H | 1702 | hCoV-19/Canada/QC-LSPQ-L00219340/2020\|EPI_ISL_5357 | ···················· | ···················· | ···················· | ···················· | ···················· | ···················· |
| H | 1703 | hCoV-19/Canada/QC-LSPQ-L00219533/2020\|EPI_ISL_5357 | ···················· | ···················· | ···················· | ···················· | ···················· | ···················· |
| H | 1704 | hCoV-19/Canada/QC-LSPQ-L00219618/2020\|EPI_ISL_5357 | ···················· | ···················· | ···················· | ···················· | ···················· | ···················· |
| H | 1705 | hCoV-19/Canada/QC-LSPQ-L00219619/2020\|EPI_ISL_5357 | ···················· | ···················· | ···················· | ···················· | ···················· | ···················· |
| H | 1706 | hCoV-19/Canada/QC-LSPQ-L00219966/2020\|EPI_ISL_5357 | ···················· | ···················· | ···················· | ···················· | ···················· | ···················· |
| H | 1707 | hCoV-19/Canada/QC-LSPQ-L00220289/2020\|EPI_ISL_5357 | ···················· | ···················· | ···················· | ···················· | ···················· | ···················· |
| H | 1708 | hCoV-19/Canada/QC-LSPQ-L00220369/2020\|EPI_ISL_5357 | ···················· | ···················· | ···················· | ···················· | ···················· | ···················· |
| H | 1709 | hCoV-19/Canada/QC-LSPQ-L00220546/2020\|EPI_ISL_5357 | ···················· | ···················· | ···················· | ···················· | ···················· | ···················· |
| H | 1716 | hCoV-19/Spain/AN-IBV-98001687/2020\|EPI_ISL_538127\| | ···················· | ···················· | ···················· | ···················· | ···················· | ···················· |
| H | 1717 | hCoV-19/Spain/AN-IBV-98001689/2020\|EPI_ISL_538128\| | ···················· | ···················· | ···················· | ···················· | ···················· | ···················· |
| H | 1718 | hCoV-19/Spain/AN-IBV-98001690/2020\|EPI_ISL_538129\| | ···················· | ···················· | ···················· | ···················· | ···················· | ···················· |
| H | 1719 | hCoV-19/Switzerland/ZH-ETHZ-260123/2020\|EPI_ISL_53 | ···················· | ···················· | ···················· | ···················· | ···················· | ···················· |
| H | 1720 | hCoV-19/Switzerland/BE-ETHZ-260125/2020\|EPI_ISL_53 | ···················· | ···················· | ···················· | ···················· | ···················· | ···················· |
| H | 1721 | hCoV-19/Switzerland/ZH-ETHZ-260126/2020\|EPI_ISL_53 | ···················· | ···················· | ···················· | ···················· | ···················· | ···················· |
| H | 1722 | hCoV-19/Switzerland/BE-ETHZ-260130/2020\|EPI_ISL_53 | ···················· | ···················· | ···················· | ···················· | ···················· | ···················· |
| H | 1723 | hCoV-19/Switzerland/AG-ETHZ-260131/2020\|EPI_ISL_53 | ···················· | ···················· | ···················· | ···················· | ···················· | ···················· |
| H | 1724 | hCoV-19/Switzerland/BE-ETHZ-260132/2020\|EPI_ISL_53 | ···················· | ···················· | ···················· | ···················· | ···················· | ···················· |
| H | 1725 | hCoV-19/Switzerland/BE-ETHZ-260133/2020\|EPI_ISL_53 | ···················· | ···················· | ···················· | ···················· | ···················· | ···················· |
| H | 1726 | hCoV-19/Switzerland/ZH-ETHZ-260143/2020\|EPI_ISL_53 | ···················· | ···················· | ···················· | ···················· | ···················· | ···················· |
| H | 1727 | hCoV-19/Switzerland/VD-ETHZ-260144/2020\|EPI_ISL_53 | ···················· | ···················· | ···················· | ···················· | ···················· | ···················· |
| H | 1728 | hCoV-19/Switzerland/VD-ETHZ-260145/2020\|EPI_ISL_53 | ···················· | ···················· | ···················· | ···················· | ···················· | ···················· |
| H | 1729 | hCoV-19/Switzerland/BE-ETHZ-260146/2020\|EPI_ISL_53 | ···················· | ···················· | ···················· | ···················· | ···················· | ···················· |
| H | 1730 | hCoV-19/Switzerland/BE-ETHZ-260147/2020\|EPI_ISL_53 | ···················· | ···················· | ···················· | ···················· | ···················· | ···················· |
| H | 1731 | hCoV-19/Switzerland/AG-ETHZ-260148/2020\|EPI_ISL_53 | ···················· | ···················· | ···················· | ···················· | ···················· | ···················· |
| H | 1732 | hCoV-19/Switzerland/AG-ETHZ-260149/2020\|EPI_ISL_53 | ···················· | ···················· | ···················· | ···················· | ···················· | ···················· |
| H | 1733 | hCoV-19/Switzerland/ZH-ETHZ-260151/2020\|EPI_ISL_53 | ···················· | ···················· | ···················· | ···················· | ···················· | ···················· |
| H | 1734 | hCoV-19/Switzerland/BE-ETHZ-260152/2020\|EPI_ISL_53 | ···················· | ···················· | ···················· | ···················· | ···················· | ···················· |
| H | 1735 | hCoV-19/Switzerland/AG-ETHZ-260153/2020\|EPI_ISL_53 | ···················· | ···················· | ···················· | ···················· | ···················· | ···················· |
| H | 1736 | hCoV-19/Switzerland/BE-ETHZ-260155/2020\|EPI_ISL_53 | ···················· | ···················· | ···················· | ···················· | ···················· | ···················· |
| H | 1737 | hCoV-19/Switzerland/AG-ETHZ-260158/2020\|EPI_ISL_53 | ···················· | ···················· | ···················· | ···················· | ···················· | ···················· |
| H | 1738 | hCoV-19/Switzerland/ZH-ETHZ-260161/2020\|EPI_ISL_53 | ···················· | ···················· | ···················· | ···················· | ···················· | ···················· |
| H | 1739 | hCoV-19/Switzerland/BL-ETHZ-260162/2020\|EPI_ISL_53 | ···················· | ·A·················· | ···················· | ···················· | ···················· | ···················· |
| H | 1740 | hCoV-19/Switzerland/BL-ETHZ-260165/2020\|EPI_ISL_53 | ···················· | ···················· | ···················· | ···················· | ·····K·············· | ···················· |
| H | 1741 | hCoV-19/Switzerland/BE-ETHZ-260166/2020\|EPI_ISL_53 | ···················· | ···················· | ···················· | ···················· | ···················· | ···················· |
| H | 1742 | hCoV-19/Switzerland/BE-ETHZ-260167/2020\|EPI_ISL_53 | ···················· | ···················· | ···················· | ···················· | ·····K·············· | ···················· |
| H | 1743 | hCoV-19/England/QEUH-96F68C/2020\|EPI_ISL_540289\|20 | ···················· | ···················· | ···················· | ···················· | ···················· | ···················· |
| H | 1744 | hCoV-19/England/QEUH-96FABD/2020\|EPI_ISL_540290\|20 | ···················· | ···················· | ···················· | ···················· | ···················· | ···················· |
| H | 1745 | hCoV-19/England/QEUH-96F8FF/2020\|EPI_ISL_540291\|20 | ···················· | ···················· | ···················· | ···················· | ···················· | ···················· |
| H | 1746 | hCoV-19/Northern Ireland/QEUH-96FDB4/2020\|EPI_ISL_ | ···················· | ···················· | ···················· | ···················· | ···················· | ···················· |
| H | 1747 | hCoV-19/Northern Ireland/QEUH-96FD1E/2020\|EPI_ISL_ | ···················· | ···················· | ···················· | ···················· | ···················· | ···················· |
| H | 1748 | hCoV-19/England/QEUH-96FF90/2020\|EPI_ISL_540294\|20 | ···················· | ···················· | ···················· | ···················· | ···················· | ···················· |
| H | 1749 | hCoV-19/Northern Ireland/QEUH-96FBBA/2020\|EPI_ISL_ | ···················· | ···················· | ···················· | ···················· | ···················· | ···················· |
| H | 1750 | hCoV-19/England/QEUH-96FD5A/2020\|EPI_ISL_540297\|20 | ···················· | ···················· | ···················· | ···················· | ···················· | ···················· |
| H | 1751 | hCoV-19/England/QEUH-96F7B6/2020\|EPI_ISL_540298\|20 | ···················· | ···················· | ···················· | ···················· | ···················· | ···················· |
| H | 1752 | hCoV-19/Northern Ireland/QEUH-97006C/2020\|EPI_ISL_ | ···················· | ···················· | ···················· | ···················· | ···················· | ···················· |
| H | 1753 | hCoV-19/England/QEUH-98EE16/2020\|EPI_ISL_540410\|20 | ···················· | ·A·················· | ···················· | ···················· | ···················· | ···················· |
| H | 1754 | hCoV-19/England/QEUH-98EFAA/2020\|EPI_ISL_540411\|20 | ···················· | ···················· | ···················· | ···················· | ···T················ | ···················· |
| H | 1755 | hCoV-19/Scotland/QEUH-98F6B3/2020\|EPI_ISL_540413\|2 | ···················· | ···················· | ···················· | ···················· | ···················· | ···················· |
| H | 1756 | hCoV-19/England/QEUH-96B055/2020\|EPI_ISL_540420\|20 | ···················· | ···················· | ···················· | ···················· | ···················· | ···················· |
| H | 1757 | hCoV-19/USA/WY-WYPHL-20011618/2020\|EPI_ISL_540421\| | ···················· | ···················· | ···················· | ···················· | ···················· | ···················· |
| H | 1758 | hCoV-19/USA/WY-WYPHL-20011656/2020\|EPI_ISL_540422\| | ···················· | ···················· | ···················· | ···················· | ···················· | ···················· |
| H | 1759 | hCoV-19/USA/IN-CDC-0821/2020\|EPI_ISL_540431\|2020-0 | ···················· | ···················· | ···················· | ···················· | ···················· | ···················· |
| H | 1760 | hCoV-19/USA/TN-CDC-0815/2020\|EPI_ISL_540432\|2020-0 | ···················· | ···················· | ···················· | ···················· | ···················· | ···················· |
| H | 1761 | hCoV-19/USA/TN-CDC-0818/2020\|EPI_ISL_540433\|2020-0 | ···················· | ···················· | ···················· | ···················· | ···················· | ···················· |
| H | 1762 | hCoV-19/USA/TN-CDC-7936/2020\|EPI_ISL_540434\|2020-0 | ···················· | ···················· | ···················· | ···················· | ···················· | ···················· |

| | | | | | | | | |
|---|---|---|---|---|---|---|---|---|
| H | 1763 | hCoV-19/USA/VA-CDC-8005/2020\|EPI_ISL_540435\|2020-0 | ···················· | ···················· | ···················· | ···················· | ···················· | ···················· |
| H | 1764 | hCoV-19/USA/WI-CDC-8093/2020\|EPI_ISL_540436\|2020-0 | ···················· | ···················· | ···················· | ···················· | ···················· | ···················· |
| H | 1765 | hCoV-19/USA/WI-CDC-8099/2020\|EPI_ISL_540437\|2020-0 | ···················· | ···················· | ···················· | ···················· | ···················· | ···················· |
| H | 1766 | hCoV-19/USA/WI-CDC-8101/2020\|EPI_ISL_540438\|2020-0 | ···················· | ···················· | ···················· | ···················· | ···················· | ···················· |
| H | 1767 | hCoV-19/USA/WV-CDC-7935/2020\|EPI_ISL_540439\|2020-0 | ···················· | ···················· | ···················· | ···················· | ···················· | ···················· |
| H | 1768 | hCoV-19/Belgium/ULG-10275/2020\|EPI_ISL_540442\|2020 | ···················· | ···················· | ···················· | ···················· | ···················· | ···················· |
| H | 1769 | hCoV-19/India/MH-NIV-76773/2020\|EPI_ISL_541741\|202 | ···················· | ···················· | ···················· | ···················· | ···················· | ···················· |
| H | 1770 | hCoV-19/India/MH-NIV-77268/2020\|EPI_ISL_541742\|202 | ···················· | ···················· | ···················· | ···················· | ···················· | ···················· |
| H | 1771 | hCoV-19/India/MH-NIV-77702/2020\|EPI_ISL_541743\|202 | ···················· | ···················· | ···················· | ···················· | ···················· | ···················· |
| H | 1772 | hCoV-19/India/MH-NIV-99913/2020\|EPI_ISL_541751\|202 | ···················· | ···················· | ···················· | ···················· | ···················· | ···················· |
| H | 1773 | hCoV-19/England/LOND-1286D60/2020\|EPI_ISL_541752\|2 | ···················· | ···················· | ···················· | ···················· | ···················· | ···················· |
| H | 1774 | hCoV-19/England/LOND-1286E03/2020\|EPI_ISL_541754\|2 | ···················· | ···················· | ···················· | ···················· | ···················· | ···················· |
| H | 1775 | hCoV-19/England/LOND-1286E12/2020\|EPI_ISL_541755\|2 | ···················· | ···················· | ···················· | ···················· | ···················· | ···················· |
| H | 1776 | hCoV-19/England/BHRT-24B4177/2020\|EPI_ISL_541756\|2 | ···················· | ···················· | ···················· | ···················· | ···················· | ···················· |
| H | 1777 | hCoV-19/England/LOND-1286E6D/2020\|EPI_ISL_541757\|2 | ···················· | ···················· | ···················· | ···················· | ···················· | ···················· |
| H | 1778 | hCoV-19/England/LOND-1286C54/2020\|EPI_ISL_541759\|2 | ···················· | ···················· | ···················· | ···················· | ···················· | ···················· |
| H | 1779 | hCoV-19/England/LOND-1286E30/2020\|EPI_ISL_541760\|2 | ···················· | ···················· | ···················· | ···················· | ···················· | ···················· |
| H | 1780 | hCoV-19/England/LOND-1286E7C/2020\|EPI_ISL_541761\|2 | ···················· | ···················· | ···················· | ···················· | ···················· | ···················· |
| H | 1781 | hCoV-19/England/LOND-1286F2E/2020\|EPI_ISL_541762\|2 | ···················· | ···················· | ···················· | ···················· | ···················· | ···················· |
| H | 1782 | hCoV-19/England/LOND-1286D9D/2020\|EPI_ISL_541763\|2 | ···················· | ····A··············· | ···················· | ···················· | ···················· | ···················· |
| H | 1783 | hCoV-19/England/LOND-1286F1F/2020\|EPI_ISL_541764\|2 | ···················· | ···················· | ···················· | ···················· | ···················· | ···················· |
| H | 1784 | hCoV-19/England/LOND-1286F3D/2020\|EPI_ISL_541765\|2 | ···················· | ···················· | ···················· | ···················· | ···················· | ···················· |
| H | 1785 | hCoV-19/England/LOND-1286F00/2020\|EPI_ISL_541766\|2 | ···················· | ···················· | ···················· | ···················· | ···················· | ···················· |
| H | 1786 | hCoV-19/England/BHRT-24B3FAB/2020\|EPI_ISL_541767\|2 | ···················· | ·A·················· | ···················· | ···················· | ···················· | ···················· |
| H | 1787 | hCoV-19/England/LOND-1286DD9/2020\|EPI_ISL_541768\|2 | ···················· | ···················· | ···················· | ···················· | ···················· | ···················· |
| H | 1788 | hCoV-19/England/LOND-1286EB8/2020\|EPI_ISL_541769\|2 | ···················· | ···················· | ···················· | ···················· | ···················· | ···················· |
| H | 1789 | hCoV-19/England/LOND-1286DE8/2020\|EPI_ISL_541770\|2 | ···················· | ···················· | ···················· | ···················· | ···················· | ···················· |
| H | 1790 | hCoV-19/England/LOND-1286ED6/2020\|EPI_ISL_541771\|2 | ···················· | ···················· | ···················· | ···················· | ···················· | ···················· |
| H | 1791 | hCoV-19/England/LOND-1286E21/2020\|EPI_ISL_541772\|2 | ···················· | ···················· | ···················· | ···················· | ···················· | ···················· |
| H | 1792 | hCoV-19/England/BHRT-24B3E17/2020\|EPI_ISL_541773\|2 | ···················· | ···················· | ···················· | ···················· | ···················· | ···················· |
| H | 1793 | hCoV-19/England/QEUH-7A7A24/2020\|EPI_ISL_533100\|20 | ···················· | ···················· | ···················· | ···················· | ···················· | ···················· |
| H | 1794 | hCoV-19/Northern Ireland/QEUH-781BF0/2020\|EPI_ISL_ | ···················· | ···················· | ···················· | ···················· | ···················· | ···················· |
| H | 1795 | hCoV-19/England/QEUH-79C4F7/2020\|EPI_ISL_533102\|20 | ···················· | ···················· | ···················· | ···················· | ···················· | ···················· |
| H | 1796 | hCoV-19/England/QEUH-796387/2020\|EPI_ISL_533103\|20 | ···················· | ···················· | ···················· | ···················· | ···················· | ···················· |
| H | 1797 | hCoV-19/England/QEUH-7A1EC4/2020\|EPI_ISL_533104\|20 | ···················· | ···················· | ···················· | ···················· | ···················· | ···················· |
| H | 1798 | hCoV-19/England/QEUH-79AE46/2020\|EPI_ISL_533105\|20 | ···················· | ·A·················· | ···················· | ···················· | ···················· | ···················· |
| H | 1799 | hCoV-19/England/QEUH-7948A8/2020\|EPI_ISL_533106\|20 | ···················· | ···················· | ···················· | ···················· | ···················· | ···················· |
| H | 1800 | hCoV-19/England/QEUH-7A0C25/2020\|EPI_ISL_533108\|20 | ···················· | ···················· | ···················· | ···················· | ···················· | ···················· |
| H | 1801 | hCoV-19/England/QEUH-788C50/2020\|EPI_ISL_533109\|20 | ···················· | ···················· | ···················· | ···················· | ···················· | ···················· |
| H | 1802 | hCoV-19/England/QEUH-79824C/2020\|EPI_ISL_533110\|20 | ···················· | ···················· | ···················· | ···················· | ···················· | ···················· |
| H | 1803 | hCoV-19/England/QEUH-769754/2020\|EPI_ISL_533111\|20 | ···················· | ···················· | ···················· | ···················· | ···················· | ···················· |
| H | 1804 | hCoV-19/England/QEUH-785181/2020\|EPI_ISL_533112\|20 | ···················· | ···················· | ···················· | ···················· | ···················· | ···················· |
| H | 1805 | hCoV-19/England/QEUH-7A774B/2020\|EPI_ISL_533113\|20 | ···················· | ···················· | ···················· | ···················· | ···················· | ···················· |
| H | 1806 | hCoV-19/England/QEUH-7A8540/2020\|EPI_ISL_533114\|20 | ···················· | ·A·················· | ···················· | ···················· | ···················· | ···A················ |
| H | 1807 | hCoV-19/England/QEUH-76BE6C/2020\|EPI_ISL_533115\|20 | ···················· | ···················· | ···················· | ···················· | ···················· | ···················· |
| H | 1808 | hCoV-19/England/QEUH-79089D/2020\|EPI_ISL_533116\|20 | ···················· | ···················· | ···················· | ···················· | ···················· | ···················· |
| H | 1809 | hCoV-19/Scotland/QEUH-964010/2020\|EPI_ISL_534401\|2 | ···················· | ···················· | ···················· | ···················· | ···················· | ···················· |
| H | 1810 | hCoV-19/Scotland/QEUH-9650F1/2020\|EPI_ISL_534402\|2 | ···················· | ···················· | ···················· | ···················· | ···················· | ···················· |
| H | 1811 | hCoV-19/Scotland/QEUH-96A1BD/2020\|EPI_ISL_534404\|2 | ···················· | ···················· | ···················· | ···················· | ···················· | ···················· |
| H | 1812 | hCoV-19/Scotland/QEUH-963AE6/2020\|EPI_ISL_534405\|2 | ···················· | ···················· | ···················· | ···················· | ···················· | ···················· |
| H | 1813 | hCoV-19/Scotland/QEUH-961E37/2020\|EPI_ISL_534406\|2 | ···················· | ···················· | ···················· | ···················· | ···················· | ···················· |
| H | 1814 | hCoV-19/Scotland/QEUH-95E152/2020\|EPI_ISL_534407\|2 | ···················· | ···················· | ···················· | ···················· | ···················· | ···················· |
| H | 1815 | hCoV-19/Scotland/QEUH-95E32F/2020\|EPI_ISL_534408\|2 | ···················· | ·A·················· | ···················· | ···················· | ···················· | ···················· |
| H | 1816 | hCoV-19/Scotland/QEUH-95E082/2020\|EPI_ISL_534409\|2 | ···················· | ···················· | ···················· | ···················· | ···················· | ···················· |
| H | 1817 | hCoV-19/Scotland/QEUH-95E459/2020\|EPI_ISL_534410\|2 | ···················· | ···················· | ···················· | ···················· | ···················· | ···················· |
| H | 1818 | hCoV-19/Scotland/QEUH-95E44A/2020\|EPI_ISL_534411\|2 | ···················· | ···················· | ···················· | ···················· | ···················· | ···················· |
| H | 1819 | hCoV-19/Scotland/QEUH-95E35C/2020\|EPI_ISL_534412\|2 | ···················· | ···················· | ···················· | ···················· | ···················· | ···················· |
| H | 1820 | hCoV-19/Scotland/QEUH-95E495/2020\|EPI_ISL_534413\|2 | ···················· | ···················· | ···················· | ···················· | ···················· | ···················· |
| H | 1821 | hCoV-19/Scotland/QEUH-95E468/2020\|EPI_ISL_534414\|2 | ···················· | ···················· | ···················· | ···················· | ···················· | ···················· |
| H | 1822 | hCoV-19/Scotland/QEUH-95E19E/2020\|EPI_ISL_534415\|2 | ···················· | ···················· | ···················· | ···················· | ···················· | ···················· |
| H | 1823 | hCoV-19/Scotland/QEUH-95E161/2020\|EPI_ISL_534418\|2 | ···················· | ···················· | ···················· | ···················· | ···················· | ···················· |
| H | 1824 | hCoV-19/Scotland/QEUH-961BB8/2020\|EPI_ISL_534419\|2 | ···················· | ···················· | ···················· | ···················· | ···················· | ···················· |
| H | 1825 | hCoV-19/Scotland/QEUH-95E1AD/2020\|EPI_ISL_534420\|2 | ···················· | ···················· | ···················· | ···················· | ···················· | ···················· |
| H | 1826 | hCoV-19/Scotland/QEUH-95DFA4/2020\|EPI_ISL_534421\|2 | ···················· | ···················· | ···················· | ···················· | ···················· | ···················· |
| H | 1827 | hCoV-19/Scotland/QEUH-95E143/2020\|EPI_ISL_534422\|2 | ···················· | ···················· | ···················· | ···················· | ···················· | ···················· |
| H | 1828 | hCoV-19/Scotland/QEUH-95E477/2020\|EPI_ISL_534423\|2 | ···················· | ···················· | ···················· | ···················· | ···················· | ···················· |
| H | 1829 | hCoV-19/Scotland/QEUH-95E486/2020\|EPI_ISL_534424\|2 | ···················· | ···················· | ···················· | ···················· | ···················· | ···················· |
| H | 1830 | hCoV-19/Scotland/QEUH-95E27D/2020\|EPI_ISL_534425\|2 | ···················· | ···················· | ···················· | ···················· | ···················· | ···················· |
| H | 1831 | hCoV-19/Scotland/QEUH-95DF95/2020\|EPI_ISL_534426\|2 | ···················· | ···················· | ···················· | ···················· | ···················· | ···················· |
| H | 1832 | hCoV-19/Scotland/QEUH-95E1DA/2020\|EPI_ISL_534427\|2 | ···················· | ·A·················· | ···················· | ···················· | ···················· | ···················· |
| H | 1833 | hCoV-19/Scotland/QEUH-95E4FF/2020\|EPI_ISL_534428\|2 | ···················· | ···················· | ···················· | ···················· | ···················· | ···················· |
| H | 1834 | hCoV-19/Scotland/QEUH-95E4C2/2020\|EPI_ISL_534429\|2 | ···················· | ···················· | ···················· | ···················· | ···················· | ···················· |
| H | 1835 | hCoV-19/Scotland/QEUH-95E3F2/2020\|EPI_ISL_534430\|2 | ···················· | ·A·················· | ···················· | ···················· | ···················· | ···················· |
| H | 1836 | hCoV-19/Scotland/QEUH-95E18F/2020\|EPI_ISL_534431\|2 | ···················· | ···················· | ···················· | ···················· | ···················· | ···················· |
| H | 1837 | hCoV-19/Scotland/QEUH-95E2AA/2020\|EPI_ISL_534433\|2 | ···················· | ···················· | ···················· | ···················· | ···················· | ···················· |
| H | 1838 | hCoV-19/Scotland/QEUH-95E26E/2020\|EPI_ISL_534434\|2 | ···················· | ···················· | ···················· | ···················· | ···················· | ···················· |
| H | 1839 | hCoV-19/Scotland/QEUH-95E4A4/2020\|EPI_ISL_534435\|2 | ···················· | ·A·················· | ···················· | ···················· | ···················· | ···················· |
| H | 1840 | hCoV-19/Scotland/QEUH-95E1F8/2020\|EPI_ISL_534436\|2 | ···················· | ···················· | ···················· | ···················· | ···················· | ···················· |
| H | 1841 | hCoV-19/Scotland/QEUH-95E091/2020\|EPI_ISL_534437\|2 | ···················· | ···················· | ···················· | ···················· | ···················· | ···················· |
| H | 1842 | hCoV-19/Scotland/QEUH-95E310/2020\|EPI_ISL_534438\|2 | ···················· | ·A·················· | ···················· | ···················· | ···················· | ···················· |

| | | | | | | | | |
|---|---|---|---|---|---|---|---|---|
| H | 1843 | hCoV-19/Scotland/QEUH-99821D/2020\|EPI_ISL_534439\|2 | ···················· | ···················· | ····················· | ···················· | ···················· | ···················· |
| H | 1844 | hCoV-19/Scotland/QEUH-99822C/2020\|EPI_ISL_534440\|2 | ···················· | ···················· | ····················· | ···················· | ···················· | ···················· |
| H | 1845 | hCoV-19/Scotland/QEUH-997FEA/2020\|EPI_ISL_534441\|2 | ···················· | ···················· | ····················· | ···················· | ···················· | ···················· |
| H | 1846 | hCoV-19/Scotland/QEUH-998259/2020\|EPI_ISL_534442\|2 | ···················· | ···················· | ····················· | ···················· | ···················· | ···················· |
| H | 1847 | hCoV-19/Scotland/QEUH-998268/2020\|EPI_ISL_534443\|2 | ···················· | ···················· | ····················· | ···················· | ···················· | ···················· |
| H | 1848 | hCoV-19/Scotland/QEUH-99820E/2020\|EPI_ISL_534444\|2 | ···················· | ···················· | ····················· | ···················· | ···················· | ···················· |
| H | 1849 | hCoV-19/Scotland/QEUH-9905A3/2020\|EPI_ISL_534445\|2 | ···················· | ·A·················· | ····················· | ···················· | ···················· | ··A················· |
| H | 1850 | hCoV-19/Scotland/QEUH-990585/2020\|EPI_ISL_534446\|2 | ···················· | ···················· | ····················· | ···················· | ···················· | ···················· |
| H | 1851 | hCoV-19/Scotland/QEUH-98FAB7/2020\|EPI_ISL_534447\|2 | ···················· | ···················· | ····················· | ···················· | ···················· | ···················· |
| H | 1852 | hCoV-19/Canada/QC-LSPQ-L00215022/2020\|EPI_ISL_5357 | ···················· | ···················· | ····················· | ···················· | ···················· | ···················· |
| H | 1853 | hCoV-19/Canada/QC-LSPQ-L00215586/2020\|EPI_ISL_5357 | ···················· | ···················· | ····················· | ···················· | ···················· | ···················· |
| H | 1854 | hCoV-19/Canada/QC-LSPQ-L00215628/2020\|EPI_ISL_5357 | ···················· | ···················· | ····················· | ···················· | ···················· | ···················· |
| H | 1855 | hCoV-19/Canada/QC-LSPQ-L00215711/2020\|EPI_ISL_5357 | ···················· | ···················· | ····················· | ···················· | ···················· | ···················· |
| H | 1856 | hCoV-19/Canada/QC-LSPQ-L00215713/2020\|EPI_ISL_5357 | ···················· | ···················· | ····················· | ···················· | ···················· | ···················· |
| H | 1857 | hCoV-19/Canada/QC-LSPQ-L00215798/2020\|EPI_ISL_5357 | ···················· | ···················· | ····················· | ···················· | ···················· | ···················· |
| H | 1858 | hCoV-19/Canada/QC-LSPQ-L00215812/2020\|EPI_ISL_5357 | ···················· | ···················· | ····················· | ···················· | ···················· | ···················· |
| H | 1859 | hCoV-19/Canada/QC-LSPQ-L00215817/2020\|EPI_ISL_5357 | ···················· | ···················· | ····················· | ···················· | ···················· | ···················· |
| H | 1860 | hCoV-19/Canada/QC-LSPQ-L00215858/2020\|EPI_ISL_5357 | ···················· | ···················· | ····················· | ···················· | ···················· | ···················· |
| H | 1861 | hCoV-19/Canada/QC-LSPQ-L00215879/2020\|EPI_ISL_5357 | ···················· | ···················· | ····················· | ···················· | ···················· | ···················· |
| H | 1862 | hCoV-19/Canada/QC-LSPQ-L00215977/2020\|EPI_ISL_5357 | ···················· | ···················· | ····················· | ···················· | ···················· | ···················· |
| H | 1863 | hCoV-19/Canada/QC-LSPQ-L00215981/2020\|EPI_ISL_5357 | ···················· | ···················· | ····················· | ···················· | ···················· | ···················· |
| H | 1864 | hCoV-19/Canada/QC-LSPQ-L00216190/2020\|EPI_ISL_5357 | ···················· | ···················· | ····················· | ···················· | ···················· | ···················· |
| H | 1865 | hCoV-19/Canada/QC-LSPQ-L00216197/2020\|EPI_ISL_5357 | ···················· | ···················· | ····················· | ···················· | ···················· | ···················· |
| H | 1866 | hCoV-19/Canada/QC-LSPQ-L00216421/2020\|EPI_ISL_5357 | ···················· | ···················· | ····················· | ···················· | ···················· | ···················· |
| H | 1867 | hCoV-19/Canada/QC-LSPQ-L00216432/2020\|EPI_ISL_5357 | ···················· | ···················· | ····················· | ···················· | ···················· | ···················· |
| H | 1868 | hCoV-19/Canada/QC-LSPQ-L00216478/2020\|EPI_ISL_5357 | ···················· | ···················· | ····················· | ···················· | ···················· | ···················· |
| H | 1869 | hCoV-19/Canada/QC-LSPQ-L00216503/2020\|EPI_ISL_5357 | ···················· | ···················· | ····················· | ···················· | ···················· | ···················· |
| H | 1870 | hCoV-19/Canada/QC-LSPQ-L00216532/2020\|EPI_ISL_5357 | ···················· | ···················· | ····················· | ···················· | ···················· | ···················· |
| H | 1871 | hCoV-19/Canada/QC-LSPQ-L00216731/2020\|EPI_ISL_5357 | ···················· | ···················· | ····················· | ···················· | ···················· | ···················· |
| H | 1872 | hCoV-19/Canada/QC-LSPQ-L00216909/2020\|EPI_ISL_5357 | ···················· | ···················· | ····················· | ···················· | ···················· | ···················· |
| H | 1873 | hCoV-19/Canada/QC-LSPQ-L00216920/2020\|EPI_ISL_5357 | ···················· | ···················· | ····················· | ···················· | ···················· | ···················· |
| H | 1874 | hCoV-19/Canada/QC-LSPQ-L00216993/2020\|EPI_ISL_5357 | ···················· | ···················· | ····················· | ···················· | ···················· | ···················· |
| H | 1875 | hCoV-19/Canada/QC-LSPQ-L00217024/2020\|EPI_ISL_5357 | ···················· | ···················· | ····················· | ···················· | ···················· | ···················· |
| H | 1876 | hCoV-19/Canada/QC-LSPQ-L00217032/2020\|EPI_ISL_5357 | ···················· | ···················· | ····················· | ···················· | ···················· | ···················· |
| H | 1877 | hCoV-19/Canada/QC-LSPQ-L00217092/2020\|EPI_ISL_5357 | ···················· | ···················· | ····················· | ···················· | ···················· | ···················· |
| H | 1878 | hCoV-19/Canada/QC-LSPQ-L00217098/2020\|EPI_ISL_5357 | ···················· | ···················· | ····················· | ···················· | ···················· | ···················· |
| H | 1879 | hCoV-19/Canada/QC-LSPQ-L00217411/2020\|EPI_ISL_5357 | ···················· | ···················· | ····················· | ···················· | ···················· | ···················· |
| H | 1880 | hCoV-19/Canada/QC-LSPQ-L00217527/2020\|EPI_ISL_5357 | ···················· | ···················· | ····················· | ···················· | ···················· | ···················· |
| H | 1881 | hCoV-19/Canada/QC-LSPQ-L00217534/2020\|EPI_ISL_5357 | ···················· | ···················· | ····················· | ···················· | ···················· | ···················· |
| H | 1882 | hCoV-19/Canada/QC-LSPQ-L00217559/2020\|EPI_ISL_5357 | ···················· | ···················· | ····················· | ···················· | ···················· | ···················· |
| H | 1883 | hCoV-19/Canada/QC-LSPQ-L00217574/2020\|EPI_ISL_5357 | ···················· | ···················· | ····················· | ···················· | ···················· | ···················· |
| H | 1884 | hCoV-19/Canada/QC-LSPQ-L00217586/2020\|EPI_ISL_5357 | ···················· | ···················· | ····················· | ···················· | ···················· | ···················· |
| H | 1885 | hCoV-19/Canada/QC-LSPQ-L00217831/2020\|EPI_ISL_5357 | ···················· | ···················· | ····················· | ···················· | ···················· | ···················· |
| H | 1886 | hCoV-19/Canada/QC-LSPQ-L00217833/2020\|EPI_ISL_5357 | ···················· | ···················· | ····················· | ···················· | ···················· | ···················· |
| H | 1887 | hCoV-19/Canada/QC-LSPQ-L00218011/2020\|EPI_ISL_5357 | ···················· | ···················· | ····················· | ···················· | ···················· | ···················· |
| H | 1888 | hCoV-19/Canada/QC-LSPQ-L00218056/2020\|EPI_ISL_5357 | ···················· | ···················· | ····················· | ···················· | ···················· | ···················· |
| H | 1889 | hCoV-19/Canada/QC-LSPQ-L00218090/2020\|EPI_ISL_5357 | ···················· | ···················· | ····················· | ···················· | ···················· | ···················· |
| H | 1890 | hCoV-19/Canada/QC-LSPQ-L00218268/2020\|EPI_ISL_5357 | ···················· | ···················· | ····················· | ···················· | ···················· | ···················· |
| H | 1891 | hCoV-19/Canada/QC-LSPQ-L00218269/2020\|EPI_ISL_5357 | ···················· | ···················· | ····················· | ···················· | ···················· | ···················· |
| H | 1892 | hCoV-19/Canada/QC-LSPQ-L00218308/2020\|EPI_ISL_5357 | ···················· | ···················· | ····················· | ···················· | ···················· | ···················· |
| H | 1893 | hCoV-19/Canada/QC-LSPQ-L00218310/2020\|EPI_ISL_5357 | ···················· | ···················· | ····················· | ···················· | ···················· | ···················· |
| H | 1894 | hCoV-19/Canada/QC-LSPQ-L00218334/2020\|EPI_ISL_5357 | ···················· | ···················· | ····················· | ···················· | ···················· | ···················· |
| H | 1895 | hCoV-19/Canada/QC-LSPQ-L00218337/2020\|EPI_ISL_5357 | ···················· | ···················· | ····················· | ···················· | ···················· | ···················· |
| H | 1896 | hCoV-19/Canada/QC-LSPQ-L00218350/2020\|EPI_ISL_5357 | ···················· | ···················· | ····················· | ···················· | ···················· | ···················· |
| H | 1897 | hCoV-19/Spain/VC-IBV-98003074/2020\|EPI_ISL_538012\| | ···················· | ···················· | ····················· | ···················· | ···················· | ···················· |
| H | 1902 | hCoV-19/Spain/VC-IBV-98003081/2020\|EPI_ISL_538017\| | ···················· | ···················· | ····················· | ···················· | ···················· | ···················· |
| H | 1904 | hCoV-19/Spain/VC-IBV-98003083/2020\|EPI_ISL_538019\| | ···················· | ···················· | ····················· | ···················· | ···················· | ···················· |
| H | 1908 | hCoV-19/Spain/VC-IBV-98003087/2020\|EPI_ISL_538023\| | ···················· | ···················· | ····················· | ···················· | ···················· | ···················· |
| H | 1909 | hCoV-19/Spain/VC-IBV-98003088/2020\|EPI_ISL_538024\| | ···················· | ···················· | ····················· | ···················· | ···················· | ···················· |
| H | 1910 | hCoV-19/Spain/VC-IBV-98003089/2020\|EPI_ISL_538025\| | ···················· | ···················· | ····················· | ···················· | ···················· | ···················· |
| H | 1917 | hCoV-19/Spain/VC-IBV-98003099/2020\|EPI_ISL_538035\| | ···················· | ···················· | ····················· | ···················· | ···················· | ···················· |
| H | 1921 | hCoV-19/Spain/VC-IBV-98003103/2020\|EPI_ISL_538039\| | ···················· | ···················· | ····················· | ···················· | ···················· | ···················· |
| H | 1924 | hCoV-19/Spain/VC-IBV-98003107/2020\|EPI_ISL_538043\| | ···················· | ···················· | ····················· | ···················· | ···················· | ···················· |
| H | 1925 | hCoV-19/Spain/VC-IBV-98003108/2020\|EPI_ISL_538044\| | ···················· | ···················· | ····················· | ···················· | ···················· | ···················· |
| H | 1926 | hCoV-19/Spain/VC-IBV-98003109/2020\|EPI_ISL_538045\| | ···················· | ···················· | ····················· | ···················· | ···················· | ···················· |
| H | 1932 | hCoV-19/Switzerland/ZH-ETHZ-260003/2020\|EPI_ISL_53 | ···················· | ···················· | ····················· | ···················· | ···················· | ···················· |
| H | 1933 | hCoV-19/Switzerland/SZ-ETHZ-260004/2020\|EPI_ISL_53 | ···················· | ···················· | ····················· | ···················· | ···················· | ···················· |
| H | 1934 | hCoV-19/Switzerland/ZH-ETHZ-260005/2020\|EPI_ISL_53 | ···················· | ···················· | ····················· | ···················· | ···················· | ···················· |
| H | 1935 | hCoV-19/Switzerland/ZH-ETHZ-260006/2020\|EPI_ISL_53 | ···················· | ···················· | ····················· | ···················· | ···················· | ···················· |
| H | 1936 | hCoV-19/Switzerland/ZH-ETHZ-260007/2020\|EPI_ISL_53 | ···················· | ···················· | ····················· | ···················· | ···················· | ···················· |
| H | 1937 | hCoV-19/Switzerland/ZH-ETHZ-260008/2020\|EPI_ISL_53 | ···················· | ···················· | ····················· | ···················· | ···················· | ···················· |
| H | 1938 | hCoV-19/Switzerland/BL-ETHZ-260009/2020\|EPI_ISL_53 | ···················· | ···G················ | ····················· | ···················· | ···················· | ···················· |
| H | 1939 | hCoV-19/Switzerland/ZH-ETHZ-260011/2020\|EPI_ISL_53 | ···················· | ···················· | ····················· | ···················· | ···················· | ···················· |
| H | 1940 | hCoV-19/Switzerland/AG-ETHZ-260013/2020\|EPI_ISL_53 | ···················· | ·A·················· | ····················· | ···················· | ···················· | ···················· |
| H | 1941 | hCoV-19/Switzerland/UR-ETHZ-260015/2020\|EPI_ISL_53 | ···················· | ···················· | ····················· | Y··················· | ···················· | ···················· |
| H | 1942 | hCoV-19/Switzerland/VD-ETHZ-260016/2020\|EPI_ISL_53 | ···················· | ···················· | ····················· | ···················· | ···················· | ···················· |
| H | 1943 | hCoV-19/Switzerland/UR-ETHZ-260018/2020\|EPI_ISL_53 | ···················· | ·R·················· | ····················· | ···················· | ···················· | ···················· |
| H | 1944 | hCoV-19/Switzerland/VD-ETHZ-260019/2020\|EPI_ISL_53 | ···················· | ···················· | ····················· | ···················· | ···················· | ···················· |
| H | 1945 | hCoV-19/Switzerland/BE-ETHZ-260022/2020\|EPI_ISL_53 | ···················· | ···················· | ····················· | ···················· | ···················· | ···················· |
| H | 1946 | hCoV-19/Switzerland/ZH-ETHZ-260023/2020\|EPI_ISL_53 | ···················· | ···················· | ····················· | ···················· | ···················· | ···················· |

| | | | | | | | | |
|---|---|---|---|---|---|---|---|---|
| H | 1947 hCoV-19/Switzerland/BL-ETHZ-260024/2020\|EPI_ISL_53 | ···················· | ···················· | ····················· | ···················· | ···················· | ···················· | |
| H | 1948 hCoV-19/Switzerland/BE-ETHZ-260025/2020\|EPI_ISL_53 | ···················· | ···················· | ····················· | ···················· | ···················· | ···················· | |
| H | 1949 hCoV-19/Switzerland/BE-ETHZ-260026/2020\|EPI_ISL_53 | ···················· | ···················· | ····················· | ···················· | ···················· | ···················· | |
| H | 1950 hCoV-19/Switzerland/ZH-ETHZ-260027/2020\|EPI_ISL_53 | ···················· | ···················· | ····················· | ···················· | ···················· | ···················· | |
| H | 1951 hCoV-19/Switzerland/ZH-ETHZ-260028/2020\|EPI_ISL_53 | ···················· | ···················· | ····················· | ···················· | ···················· | ···················· | |
| H | 1952 hCoV-19/Switzerland/BL-ETHZ-260031/2020\|EPI_ISL_53 | ···················· | ···················· | ····················· | ···················· | ···················· | ···················· | |
| H | 1953 hCoV-19/Switzerland/BL-ETHZ-260032/2020\|EPI_ISL_53 | ···················· | ···················· | ····················· | ···················· | ···················· | ···················· | |
| H | 1954 hCoV-19/Switzerland/BE-ETHZ-260034/2020\|EPI_ISL_53 | ···················· | ···················· | ····················· | ···················· | ···················· | ···················· | |
| H | 1955 hCoV-19/Switzerland/GR-ETHZ-260035/2020\|EPI_ISL_53 | ···················· | ···················· | ····················· | ···················· | ······K············· | ···················· | |
| H | 1956 hCoV-19/Switzerland/VD-ETHZ-260038/2020\|EPI_ISL_53 | ···················· | ···················· | ····················· | ···················· | ···················· | ···················· | |
| H | 1957 hCoV-19/Switzerland/VD-ETHZ-260039/2020\|EPI_ISL_53 | ···················· | ···················· | ····················· | ···················· | ···················· | ···················· | |
| H | 1958 hCoV-19/Switzerland/VD-ETHZ-260040/2020\|EPI_ISL_53 | ···················· | ···················· | ····················· | ···················· | ···················· | ···················· | |
| H | 1959 hCoV-19/Switzerland/VS-ETHZ-260042/2020\|EPI_ISL_53 | ···················· | ···················· | ····················· | ···················· | ···················· | ···················· | |
| H | 1960 hCoV-19/Switzerland/BE-ETHZ-260043/2020\|EPI_ISL_53 | ···················· | ···················· | ····················· | ···················· | ···················· | ···················· | |
| H | 1961 hCoV-19/Switzerland/BE-ETHZ-260045/2020\|EPI_ISL_53 | ···················· | ···················· | ····················· | ···················· | ···················· | ···················· | |
| H | 1962 hCoV-19/Switzerland/BL-ETHZ-260048/2020\|EPI_ISL_53 | ···················· | ····G··············· | ····················· | ···················· | ···················· | ···················· | |
| H | 1963 hCoV-19/Switzerland/BS-ETHZ-260049/2020\|EPI_ISL_53 | ···················· | ···················· | ····················· | ···················· | ···················· | ···················· | |
| H | 1964 hCoV-19/Switzerland/BS-ETHZ-260051/2020\|EPI_ISL_53 | ···················· | ·A·················· | ····················· | ···················· | ···················· | ···················· | |
| H | 1965 hCoV-19/Switzerland/ZH-ETHZ-260052/2020\|EPI_ISL_53 | ···················· | ···················· | ····················· | ···················· | ···················· | ···················· | |
| H | 1966 hCoV-19/Switzerland/BL-ETHZ-260054/2020\|EPI_ISL_53 | ··················Y· | ···················· | ····················· | ···················· | ···················· | ···················· | 1 |
| H | 1967 hCoV-19/England/QEUH-96FAAE/2020\|EPI_ISL_540322\|20 | ···················· | ···················· | ····················· | ···················· | ···················· | ···················· | |
| H | 1968 hCoV-19/England/QEUH-96FC7B/2020\|EPI_ISL_540323\|20 | ···················· | ···················· | ····················· | ···················· | ···················· | ···················· | |
| H | 1969 hCoV-19/Northern Ireland/QEUH-96FB8D/2020\|EPI_ISL_ | ···················· | ···················· | ····················· | ···················· | ···················· | ···················· | |
| H | 1970 hCoV-19/England/QEUH-96FB6F/2020\|EPI_ISL_540325\|20 | ···················· | ···················· | ····················· | ···················· | ···················· | ···················· | |
| H | 1971 hCoV-19/England/QEUH-96FA71/2020\|EPI_ISL_540326\|20 | ···················· | ···················· | ····················· | ···················· | ···················· | ···················· | |
| H | 1972 hCoV-19/England/QEUH-96F789/2020\|EPI_ISL_540328\|20 | ···················· | ···················· | ····················· | ···················· | ···················· | ···················· | |
| H | 1973 hCoV-19/England/QEUH-96FF18/2020\|EPI_ISL_540330\|20 | ···················· | ···················· | ····················· | ···················· | ···················· | ···················· | |
| H | 1974 hCoV-19/Scotland/QEUH-96F886/2020\|EPI_ISL_540331\|2 | ···················· | ···················· | ····················· | ···················· | ···················· | ···················· | |
| H | 1975 hCoV-19/Northern Ireland/QEUH-96FDA5/2020\|EPI_ISL_ | ···················· | ···················· | ····················· | ···················· | ···················· | ···················· | |
| H | 1976 hCoV-19/Scotland/QEUH-96F613/2020\|EPI_ISL_540336\|2 | ···················· | ···················· | ····················· | ···················· | ···················· | ···················· | |
| H | 1977 hCoV-19/England/QEUH-96F81D/2020\|EPI_ISL_540337\|20 | ···················· | ···················· | ····················· | ···················· | ···················· | ···················· | |
| H | 1978 hCoV-19/Northern Ireland/QEUH-96FCF3/2020\|EPI_ISL_ | ···················· | ···················· | ····················· | ···················· | ···················· | ···················· | |
| H | 1979 hCoV-19/England/QEUH-96FC11/2020\|EPI_ISL_540339\|20 | ···················· | ···················· | ····················· | ···················· | ···················· | ···················· | |
| H | 1980 hCoV-19/England/QEUH-96F877/2020\|EPI_ISL_540340\|20 | ···················· | ···················· | ····················· | ···················· | ···················· | ···················· | |
| H | 1981 hCoV-19/England/QEUH-96F65F/2020\|EPI_ISL_540341\|20 | ···················· | ···················· | ····················· | ···················· | ···················· | ···················· | |
| H | 1982 hCoV-19/England/QEUH-98F1A3/2020\|EPI_ISL_540342\|20 | ···················· | ···················· | ····················· | ···················· | ···················· | ···················· | |
| H | 1983 hCoV-19/Scotland/QEUH-98F413/2020\|EPI_ISL_540343\|2 | ···················· | ···················· | ····················· | ···················· | ···················· | ···················· | |
| H | 1984 hCoV-19/England/QEUH-98F246/2020\|EPI_ISL_540344\|20 | ···················· | ···················· | ····················· | ···················· | ···················· | ···················· | |
| H | 1985 hCoV-19/England/QEUH-98EE34/2020\|EPI_ISL_540345\|20 | ···················· | ·A·················· | ····················· | ···················· | ···················· | ···················· | |
| H | 1986 hCoV-19/England/QEUH-98F2CE/2020\|EPI_ISL_540346\|20 | ···················· | ···················· | ····················· | ···················· | ···················· | ···················· | |
| H | 1987 hCoV-19/England/QEUH-98EFB9/2020\|EPI_ISL_540347\|20 | ···················· | ···················· | ····················· | ···················· | ···················· | ···················· | |
| H | 1988 hCoV-19/England/QEUH-98F0B5/2020\|EPI_ISL_540348\|20 | ···················· | ···················· | ····················· | ···················· | ···················· | ···················· | |
| H | 1989 hCoV-19/Scotland/QEUH-98F3BC/2020\|EPI_ISL_540349\|2 | ···················· | ···················· | ····················· | ···················· | ···················· | ···················· | |
| H | 1990 hCoV-19/England/QEUH-98EF7D/2020\|EPI_ISL_540350\|20 | ···················· | ·A·················· | ····················· | ···················· | ···················· | ···················· | |
| H | 1991 hCoV-19/England/QEUH-98F1EF/2020\|EPI_ISL_540352\|20 | ···················· | ···················· | ····················· | ···················· | ···················· | ···················· | |
| H | 1992 hCoV-19/England/QEUH-98EF9B/2020\|EPI_ISL_540353\|20 | ···················· | ·A·················· | ····················· | ···················· | ···················· | ···················· | |
| H | 1993 hCoV-19/England/QEUH-98EEAD/2020\|EPI_ISL_540354\|20 | ···················· | ···················· | ····················· | ···················· | ···················· | ···················· | |
| H | 1994 hCoV-19/Scotland/QEUH-98F52F/2020\|EPI_ISL_540355\|2 | ···················· | ···················· | ····················· | ···················· | ···················· | ···················· | |
| H | 1995 hCoV-19/Scotland/QEUH-98F6A4/2020\|EPI_ISL_540356\|2 | ···················· | ···················· | ····················· | ···················· | ···················· | ···················· | |
| H | 1996 hCoV-19/England/QEUH-98F2DD/2020\|EPI_ISL_540357\|20 | ···················· | ···················· | ····················· | ···················· | ···················· | ···················· | |
| H | 1997 hCoV-19/England/QEUH-98EF5F/2020\|EPI_ISL_540358\|20 | ···················· | ···················· | ····················· | ···················· | ···················· | ···················· | |
| H | 1998 hCoV-19/England/QEUH-98EF40/2020\|EPI_ISL_540359\|20 | ···················· | ···················· | ····················· | ···················· | ···················· | ···················· | |
| H | 1999 hCoV-19/England/QEUH-98F05B/2020\|EPI_ISL_540360\|20 | ···················· | ···················· | ····················· | ···················· | ···················· | ···················· | |
| H | 2000 hCoV-19/England/QEUH-98EEBC/2020\|EPI_ISL_540361\|20 | ···················· | ···················· | ····················· | ···················· | ···················· | ···················· | |
| H | 2001 hCoV-19/Scotland/QEUH-98F404/2020\|EPI_ISL_540362\|2 | ···················· | ···················· | ····················· | ···················· | ···················· | ···················· | |
| H | 2002 hCoV-19/England/QEUH-98F4E6/2020\|EPI_ISL_540363\|20 | ···················· | ···················· | ····················· | ···················· | ···················· | ···················· | |
| H | 2003 hCoV-19/England/QEUH-98F237/2020\|EPI_ISL_540364\|20 | ···················· | ·A·················· | ····················· | ···················· | ···················· | ···A················ | |
| H | 2004 hCoV-19/Scotland/QEUH-98F510/2020\|EPI_ISL_540365\|2 | ···················· | ···················· | ····················· | ···················· | ···················· | ···················· | |
| H | 2005 hCoV-19/Serbia/NP-0332007/2020\|EPI_ISL_541653\|2020 | ···················· | ···················· | ····················· | ···················· | ···················· | ···················· | |
| H | 2006 hCoV-19/Serbia/NP-0432207/2020\|EPI_ISL_541654\|2020 | ···················· | ···················· | ····················· | ···················· | ···················· | ···················· | |
| H | 2007 hCoV-19/Serbia/KM-09962407/2020\|EPI_ISL_541655\|202 | ···················· | ···················· | ····················· | ···················· | ···················· | ···················· | |
| H | 2008 hCoV-19/Serbia/KV-0140707/2020\|EPI_ISL_541656\|2020 | ···················· | ···················· | ····················· | ···················· | ···················· | ···················· | |
| H | 2009 hCoV-19/Serbia/KM-0560707/2020\|EPI_ISL_541657\|2020 | ···················· | ···················· | ····················· | ···················· | ···················· | ···················· | |
| H | 2010 hCoV-19/Serbia/KM-050707/2020\|EPI_ISL_541658\|2020- | ···················· | ···················· | ····················· | ···················· | ···················· | ···················· | |
| H | 2011 hCoV-19/Serbia/NP-0200907/2020\|EPI_ISL_541659\|2020 | ···················· | ···················· | ····················· | ···················· | ···················· | ···················· | |
| H | 2012 hCoV-19/Serbia/KV-072107/2020\|EPI_ISL_541660\|2020- | ···················· | ···················· | ····················· | ···················· | ···················· | ···················· | |
| H | 2013 hCoV-19/Serbia/NP-0541007/2020\|EPI_ISL_541661\|2020 | ···················· | ···················· | ····················· | ···················· | ···················· | ···················· | |
| H | 2014 hCoV-19/Serbia/KV-0391607/2020\|EPI_ISL_541662\|2020 | ···················· | ···················· | ····················· | ···················· | ···················· | ···················· | |
| H | 2015 hCoV-19/USA/SC-DHEC-1037/2020\|EPI_ISL_541663\|2020- | ···················· | ···················· | ····················· | ···················· | ···················· | ···················· | |
| H | 2016 hCoV-19/USA/SC-DHEC-1079/2020\|EPI_ISL_541664\|2020- | ···················· | ···················· | ····················· | ···················· | ···················· | ···················· | |
| H | 2017 hCoV-19/USA/SC-DHEC-1081/2020\|EPI_ISL_541665\|2020- | ···················· | ···················· | ····················· | ···················· | ···················· | ···················· | |
| H | 2018 hCoV-19/USA/SC-DHEC-1083/2020\|EPI_ISL_541666\|2020- | ···················· | ···················· | ····················· | ···················· | ···················· | ···················· | |
| H | 2019 hCoV-19/USA/SC-DHEC-1084/2020\|EPI_ISL_541667\|2020- | ···················· | ·········A·········· | ····················· | ···················· | ···················· | ···················· | |
| H | 2020 hCoV-19/USA/SC-DHEC-1092/2020\|EPI_ISL_541668\|2020- | ···················· | ···················· | ····················· | ···················· | ···················· | ···················· | |
| H | 2021 hCoV-19/USA/SC-DHEC-1093/2020\|EPI_ISL_541669\|2020- | ···················· | ···················· | ····················· | ···················· | ···················· | ···················· | |
| H | 2022 hCoV-19/USA/SC-DHEC-1095/2020\|EPI_ISL_541670\|2020- | ···················· | ·A·················· | ····················· | ···················· | ···················· | ···················· | |
| H | 2023 hCoV-19/USA/SC-DHEC-0932/2020\|EPI_ISL_541672\|2020- | ···················· | ···················· | ····················· | ···················· | ···················· | ···················· | |
| H | 2024 hCoV-19/USA/SC-DHEC-0934/2020\|EPI_ISL_541673\|2020- | ···················· | ···················· | ····················· | ···················· | ···················· | ···················· | |
| H | 2025 hCoV-19/USA/SC-DHEC-0935/2020\|EPI_ISL_541674\|2020- | ···················· | ·A·················· | ····················· | ···················· | ···················· | ···················· | |
| H | 2026 hCoV-19/USA/SC-DHEC-0937/2020\|EPI_ISL_541675\|2020- | ···················· | ···················· | ····················· | ···················· | ···················· | ···················· | |

```
H    2027 hCoV-19/USA/SC-DHEC-0942/2020|EPI_ISL_541677|2020-    ····················  ····················  ·····················  ····················  ····················  ····················
H    2028 hCoV-19/USA/SC-DHEC-0943/2020|EPI_ISL_541678|2020-    ····················  ····················  ·····················  ····················  ····················  ····················
H    2029 hCoV-19/USA/SC-DHEC-0945/2020|EPI_ISL_541679|2020-    ····················  ····················  ·····················  ····················  ····················  ····················
H    2033 hCoV-19/India/MH-NIV-112247/2020|EPI_ISL_541684|20    ····················  ····················  ·····················  ····················  ····················  ····················
H    2035 hCoV-19/India/MH-NIV-116690/2020|EPI_ISL_541687|20    ····················  ····················  ·····················  ····················  ····················  ····················
H    2036 hCoV-19/India/MH-NIV-119994/2020|EPI_ISL_541688|20    ····················  ····················  ·····················  ····················  ····················  ····················
H    2038 hCoV-19/India/MH-NIV-122530/2020|EPI_ISL_541690|20    ····················  ····················  ·····················  ····················  ····················  ····················
H    2040 hCoV-19/India/MH-NIV-124593/2020|EPI_ISL_541692|20    ····················  ····················  ·····················  ·······N············  ····················  ····················
H    2041 hCoV-19/India/MH-NIV-125643/2020|EPI_ISL_541693|20    ····················  ····················  ·····················  ····················  ····················  ····················
H    2042 hCoV-19/India/MH-NIV-126959/2020|EPI_ISL_541694|20    ····················  ····················  ·····················  ····················  ····················  ····················
H    2043 hCoV-19/India/MH-NIV-136271/2020|EPI_ISL_541695|20    ····················  ····················  ·····················  ····················  ····················  ····················
H    2044 hCoV-19/India/MH-NIV-137244/2020|EPI_ISL_541696|20    ····················  ····················  ·····················  ····················  ····················  ····················
H    2045 hCoV-19/USA/NM-UNM-00570/2020|EPI_ISL_542984|2020-    ····················  ····················  ·····················  ····················  ····················  ····················
H    2046 hCoV-19/USA/NM-UNM-00574/2020|EPI_ISL_542988|2020-    ····················  ····················  ·····················  ····················  ····················  ····················
H    2047 hCoV-19/USA/NM-UNM-00575/2020|EPI_ISL_542989|2020-    ····················  ·A··················  ·····················  ····················  ····················  ····················
H    2048 hCoV-19/USA/NM-UNM-00585/2020|EPI_ISL_542997|2020-    ····················  ····················  ·····················  ····················  ····················  ····················
H    2049 hCoV-19/Northern Ireland/QEUH-96C054/2020|EPI_ISL_    ····················  ····················  ·····················  ····················  ····················  ····················
H    2050 hCoV-19/England/QEUH-96C230/2020|EPI_ISL_530922|20    ····················  ····················  ·····················  ····················  ····················  ····················
H    2051 hCoV-19/Scotland/QEUH-969FE1/2020|EPI_ISL_530923|2    ····················  ····················  ·····················  ····················  ····················  ····················
H    2052 hCoV-19/England/QEUH-96C4D0/2020|EPI_ISL_530924|20    ····················  ·A··················  ·····················  ····················  ····················  ···A················
H    2053 hCoV-19/England/QEUH-96C24F/2020|EPI_ISL_530926|20    ····················  ····················  ·····················  ····················  ····················  ····················
H    2054 hCoV-19/England/QEUH-96A8A9/2020|EPI_ISL_530927|20    ····················  ····················  ·····················  ····················  ····················  ····················
H    2055 hCoV-19/England/QEUH-96BFFE/2020|EPI_ISL_530928|20    ····················  ····················  ·····················  ····················  ····················  ····················
H    2056 hCoV-19/Scotland/QEUH-96C6CB/2020|EPI_ISL_530929|2    ····················  ····················  ·····················  ····················  ····················  ····················
H    2057 hCoV-19/England/QEUH-96C634/2020|EPI_ISL_530930|20    ····················  ····················  ·····················  ····················  ····················  ····················
H    2058 hCoV-19/England/QEUH-96C661/2020|EPI_ISL_530932|20    ····················  ·A··················  ·····················  ····················  ····················  ····················
H    2059 hCoV-19/England/QEUH-96C546/2020|EPI_ISL_530933|20    ····················  ····················  ·····················  ····················  ····················  ····················
H    2060 hCoV-19/Northern Ireland/QEUH-96C0EB/2020|EPI_ISL_    ····················  ····················  ·····················  ····················  ····················  ····················
H    2061 hCoV-19/England/QEUH-96A900/2020|EPI_ISL_530935|20    ····················  ····················  ·····················  ····················  ····················  ····················
H    2062 hCoV-19/Northern Ireland/QEUH-96C0AF/2020|EPI_ISL_    ····················  ····················  ·····················  ····················  ····················  ····················
H    2063 hCoV-19/England/QEUH-96C670/2020|EPI_ISL_530938|20    ····················  ····················  ·····················  ····················  ····················  ····················
H    2064 hCoV-19/England/QEUH-96C449/2020|EPI_ISL_530939|20    ····················  ····················  ·····················  ····················  ····················  ····················
H    2065 hCoV-19/Northern Ireland/QEUH-96C3D3/2020|EPI_ISL_    ····················  ····················  ·····················  ····················  ····················  ····················
H    2066 hCoV-19/Northern Ireland/QEUH-969E20/2020|EPI_ISL_    ····················  ····················  ·····················  ····················  ····················  ····················
H    2067 hCoV-19/England/QEUH-96C5FB/2020|EPI_ISL_530942|20    ····················  ····················  ·····················  ····················  ····················  ····················
H    2068 hCoV-19/England/QEUH-96C573/2020|EPI_ISL_530944|20    ····················  ····················  ·····················  ····················  ····················  ····················
H    2069 hCoV-19/England/QEUH-969D05/2020|EPI_ISL_530945|20    ····················  ····················  ·····················  ····················  ····················  ····················
H    2070 hCoV-19/Northern Ireland/QEUH-969DE7/2020|EPI_ISL_    ····················  ····················  ·····················  ····················  ····················  ····················
H    2071 hCoV-19/England/QEUH-96C379/2020|EPI_ISL_530947|20    ····················  ····················  ·····················  ····················  ····················  ····················
H    2072 hCoV-19/Northern Ireland/QEUH-969E5D/2020|EPI_ISL_    ····················  ····················  ·····················  ····················  ····················  ····················
H    2073 hCoV-19/England/QEUH-9696DD/2020|EPI_ISL_530949|20    ····················  ·A··················  ·····················  ····················  ····················  ···A················
H    2074 hCoV-19/England/QEUH-9652BF/2020|EPI_ISL_530950|20    ····················  ····················  ·····················  ····················  ····················  ····················
H    2075 hCoV-19/Northern Ireland/QEUH-96993E/2020|EPI_ISL_    ····················  ····················  ·····················  ····················  ····················  ····················
H    2076 hCoV-19/England/QEUH-9698D7/2020|EPI_ISL_530952|20    ····················  ····················  ·····················  ····················  ····················  ····················
H    2077 hCoV-19/Scotland/QEUH-969637/2020|EPI_ISL_530953|2    ····················  ····················  ·····················  ····················  ····················  ····················
H    2078 hCoV-19/England/QEUH-969A0E/2020|EPI_ISL_530954|20    ····················  ····················  ·····················  ····················  ····················  ····················
H    2079 hCoV-19/England/QEUH-964DEC/2020|EPI_ISL_530955|20    ····················  ····················  ·····················  ····················  ····················  ····················
H    2080 hCoV-19/Northern Ireland/QEUH-969AD1/2020|EPI_ISL_    ····················  ····················  ·····················  ····················  ····················  ····················
H    2081 hCoV-19/England/QEUH-965501/2020|EPI_ISL_530957|20    ····················  ····················  ·····················  ····················  ····················  ····················
H    2082 hCoV-19/Scotland/QEUH-969831/2020|EPI_ISL_530959|2    ····················  ····················  ·····················  ····················  ····················  ····················
H    2083 hCoV-19/England/QEUH-969479/2020|EPI_ISL_530960|20    ····················  ····················  ·····················  ····················  ····················  ····················
H    2084 hCoV-19/Scotland/QEUH-965729/2020|EPI_ISL_530961|2    ····················  ·A··················  ·····················  ····················  ····················  ····················
H    2085 hCoV-19/Scotland/QEUH-964D28/2020|EPI_ISL_530962|2    ····················  ····················  ·····················  ····················  ····················  ····················
H    2086 hCoV-19/England/QEUH-96563B/2020|EPI_ISL_530964|20    ····················  ·A··················  ·····················  ····················  ····················  ····················
H    2087 hCoV-19/England/QEUH-969619/2020|EPI_ISL_530966|20    ····················  ····················  ·····················  ····················  ····················  ····················
H    2088 hCoV-19/England/QEUH-9656C2/2020|EPI_ISL_530967|20    ····················  ····················  ·····················  ····················  ····················  ····················
H    2089 hCoV-19/England/QEUH-9657CF/2020|EPI_ISL_530968|20    ····················  ····················  ·····················  ····················  ····················  ····················
H    2090 hCoV-19/England/QEUH-9657B0/2020|EPI_ISL_530969|20    ····················  ·A··················  ·····················  ····················  ····················  ···A················
H    2091 hCoV-19/England/QEUH-96504C/2020|EPI_ISL_530970|20    ····················  ····················  ·····················  ····················  ····················  ····················
H    2092 hCoV-19/Scotland/QEUH-965774/2020|EPI_ISL_530973|2    ····················  ····················  ·····················  ····················  ····················  ····················
H    2093 hCoV-19/England/QEUH-96497F/2020|EPI_ISL_530974|20    ····················  ····················  ·····················  ····················  ····················  ····················
H    2094 hCoV-19/Scotland/QEUH-969AFF/2020|EPI_ISL_530976|2    ····················  ····················  ·····················  ····················  ····················  ····················
H    2095 hCoV-19/England/QEUH-965413/2020|EPI_ISL_530977|20    ····················  ····················  ·····················  ····················  ····················  ····················
H    2096 hCoV-19/England/QEUH-9659AB/2020|EPI_ISL_530978|20    ····················  ····················  ·····················  ····················  ····················  ····················
H    2097 hCoV-19/Scotland/QEUH-965792/2020|EPI_ISL_530979|2    ····················  ····················  ·····················  ····················  ····················  ····················
H    2098 hCoV-19/Northern Ireland/QEUH-9699A7/2020|EPI_ISL_    ····················  ····················  ·····················  ····················  ····················  ····················
H    2099 hCoV-19/England/QEUH-964D91/2020|EPI_ISL_530981|20    ····················  ····················  ·····················  ····················  ····················  ····················
H    2100 hCoV-19/Northern Ireland/QEUH-9699D4/2020|EPI_ISL_    ····················  ····················  ·····················  ····················  ····················  ····················
H    2101 hCoV-19/Scotland/QEUH-969646/2020|EPI_ISL_530983|2    ····················  ····················  ·····················  ····················  ····················  ····················
H    2102 hCoV-19/Northern Ireland/QEUH-964F7D/2020|EPI_ISL_    ····················  ····················  ·····················  ····················  ····················  ····················
H    2103 hCoV-19/Scotland/QEUH-9654B9/2020|EPI_ISL_530985|2    ····················  ·A··················  ·····················  ····················  ····················  ····················
H    2104 hCoV-19/Scotland/QEUH-964AC7/2020|EPI_ISL_530986|2    ····················  ····················  ·····················  ····················  ····················  ····················
H    2105 hCoV-19/Northern Ireland/QEUH-9699C5/2020|EPI_ISL_    ····················  ····················  ·····················  ····················  ····················  ····················
H    2106 hCoV-19/England/QEUH-96570B/2020|EPI_ISL_530989|20    ····················  ····················  ·····················  ····················  ····················  ····················
H    2107 hCoV-19/England/QEUH-964C0D/2020|EPI_ISL_530991|20    ····················  ····················  ·····················  ····················  ····················  ····················
H    2108 hCoV-19/England/QEUH-96953A/2020|EPI_ISL_530992|20    ····················  ····················  ·····················  ····················  ····················  ····················
H    2109 hCoV-19/Northern Ireland/QEUH-964DBF/2020|EPI_ISL_    ····················  ····················  ·····················  ····················  ····················  ····················
H    2110 hCoV-19/England/QEUH-9649BB/2020|EPI_ISL_530994|20    ····················  ·A··················  ·····················  ····················  ····················  ····················
H    2111 hCoV-19/England/QEUH-9658AE/2020|EPI_ISL_530995|20    ····················  ·A··················  ·····················  ····················  ····················  ···A················
H    2112 hCoV-19/Scotland/QEUH-9659BA/2020|EPI_ISL_530997|2    ····················  ····················  ·····················  ····················  ····················  ····················
```

| | | | | | | | | |
|---|---|---|---|---|---|---|---|---|
| H | 2113 | hCoV-19/England/QEUH-964FAA/2020\|EPI_ISL_530998\|20 | ···················· | ···················· | ···················· | ···················· | ···················· | ···················· |
| H | 2114 | hCoV-19/England/QEUH-964A9A/2020\|EPI_ISL_530999\|20 | ···················· | ···················· | ···················· | ···················· | ···················· | ···················· |
| H | 2115 | hCoV-19/Canada/QC-LSPQ-L00220634/2020\|EPI_ISL_5358 | ···················· | ···················· | ···················· | ···················· | ···················· | ···················· |
| H | 2116 | hCoV-19/Canada/QC-LSPQ-L00220733/2020\|EPI_ISL_5358 | ···················· | ···················· | ···················· | ···················· | ···················· | ···················· |
| H | 2117 | hCoV-19/Canada/QC-LSPQ-L00220736/2020\|EPI_ISL_5358 | ···················· | ···················· | ···················· | ···················· | ···················· | ···················· |
| H | 2118 | hCoV-19/Canada/QC-LSPQ-L00220827/2020\|EPI_ISL_5358 | ···················· | ···················· | ···················· | ···················· | ···················· | ···················· |
| H | 2119 | hCoV-19/Canada/QC-LSPQ-L00220832/2020\|EPI_ISL_5358 | ···················· | ···················· | ···················· | ···················· | ···················· | ···················· |
| H | 2120 | hCoV-19/Canada/QC-LSPQ-L00220897/2020\|EPI_ISL_5358 | ···················· | ···················· | ···················· | ···················· | ···················· | ···················· |
| H | 2121 | hCoV-19/Canada/QC-LSPQ-L00220933/2020\|EPI_ISL_5358 | ···················· | ···················· | ···················· | ···················· | ···················· | ···················· |
| H | 2122 | hCoV-19/Canada/QC-LSPQ-L00221340/2020\|EPI_ISL_5358 | ···················· | ···················· | ···················· | ···················· | ···················· | ···················· |
| H | 2123 | hCoV-19/Canada/QC-LSPQ-L00221391/2020\|EPI_ISL_5358 | ···················· | ···················· | ···················· | ···················· | ···················· | ···················· |
| H | 2124 | hCoV-19/Canada/QC-LSPQ-L00221605/2020\|EPI_ISL_5358 | ···················· | ···················· | ···················· | ···················· | ···················· | ···················· |
| H | 2125 | hCoV-19/Canada/QC-LSPQ-L00221627/2020\|EPI_ISL_5358 | ···················· | ···················· | ···················· | ···················· | ···················· | ···················· |
| H | 2126 | hCoV-19/Canada/QC-LSPQ-L00221852/2020\|EPI_ISL_5358 | ···················· | ···················· | ···················· | ···················· | ···················· | ···················· |
| H | 2127 | hCoV-19/Spain/RI-IBV-99005860/2020\|EPI_ISL_537403\| | ···················· | ···················· | ···················· | ···················· | ···················· | ···················· |
| H | 2129 | hCoV-19/Spain/RI-IBV-99005863/2020\|EPI_ISL_537406\| | ···················· | ···················· | ···················· | ···················· | ···················· | ···················· |
| H | 2130 | hCoV-19/Spain/RI-IBV-99005867/2020\|EPI_ISL_537410\| | ···················· | ···················· | ···················· | ···················· | ··N················· | ···················· |
| H | 2132 | hCoV-19/Spain/RI-IBV-99005869/2020\|EPI_ISL_537412\| | ···················· | ···················· | ···················· | ···················· | ·NN················· | ···················· |
| H | 2133 | hCoV-19/Spain/RI-IBV-99005871/2020\|EPI_ISL_537414\| | ···················· | ···················· | ···················· | ···················· | ···················· | ···················· |
| H | 2134 | hCoV-19/Spain/VC-IBV-98003115/2020\|EPI_ISL_538051\| | ···················· | ···················· | ···················· | ···················· | ···················· | ···················· |
| H | 2135 | hCoV-19/Spain/VC-IBV-98003116/2020\|EPI_ISL_538052\| | ···················· | ···················· | ···················· | ···················· | ···················· | ···················· |
| H | 2136 | hCoV-19/Spain/VC-IBV-98003118/2020\|EPI_ISL_538054\| | ···················· | ···················· | ···················· | ···················· | ···················· | ···················· |
| H | 2137 | hCoV-19/England/LEED-2A975D/2020\|EPI_ISL_538748\|20 | ···················· | ···················· | ···················· | ···················· | ···················· | ···················· |
| H | 2138 | hCoV-19/England/LEED-2A97B7/2020\|EPI_ISL_538749\|20 | ···················· | ···················· | ···················· | ···················· | ···················· | ···················· |
| H | 2139 | hCoV-19/env/Netherlands/NH-92852-N1/2020\|EPI_ISL_5 | ···················· | ···················· | ···················· | ···················· | ···················· | ···················· |
| H | 2140 | hCoV-19/Australia/NSW2754/2020\|EPI_ISL_539327\|2020 | ···················· | ···················· | ···················· | ···················· | ···················· | ···················· |
| H | 2141 | hCoV-19/Hunan/HN-CIDC-P2/2020\|EPI_ISL_539333\|2020- | ···················· | ···················· | ···················· | ···················· | ···················· | ···················· |
| H | 2142 | hCoV-19/Hunan/HN-CIDC-P5/2020\|EPI_ISL_539334\|2020- | ···················· | ···················· | ···················· | ···················· | ···················· | ···················· |
| H | 2143 | hCoV-19/Hunan/HN-CIDC-P6/2020\|EPI_ISL_539335\|2020- | ···················· | ···················· | ···················· | ···················· | ···················· | ···················· |
| H | 2144 | hCoV-19/Hunan/HN-CIDC-P7/2020\|EPI_ISL_539336\|2020- | ···················· | ···················· | ···················· | ···················· | ···················· | ···················· |
| H | 2145 | hCoV-19/Hunan/HN-CIDC-P8/2020\|EPI_ISL_539337\|2020- | ···················· | ···················· | ···················· | ···················· | ···················· | ···················· |
| H | 2146 | hCoV-19/Hunan/HN-CIDC-P10/2020\|EPI_ISL_539338\|2020 | ···················· | ···················· | ···················· | ···················· | ···················· | ···················· |
| H | 2147 | hCoV-19/Hunan/HN-CIDC-P11/2020\|EPI_ISL_539339\|2020 | ···················· | ···················· | ···················· | ···················· | ···················· | ···················· |
| H | 2148 | hCoV-19/Switzerland/ZH-ETHZ-260055/2020\|EPI_ISL_53 | ···················· | ···················· | ···················· | ···················· | ···················· | ···················· |
| H | 2149 | hCoV-19/Switzerland/AG-ETHZ-260057/2020\|EPI_ISL_53 | ···················· | ···················· | ···················· | ···················· | ···················· | ···················· |
| H | 2150 | hCoV-19/Switzerland/BL-ETHZ-260058/2020\|EPI_ISL_53 | ···················· | ···················· | ···················· | ···················· | ···················· | ···················· |
| H | 2151 | hCoV-19/Switzerland/AG-ETHZ-260060/2020\|EPI_ISL_53 | ···················· | ·A·················· | ···················· | ···················· | ···················· | ···················· |
| H | 2152 | hCoV-19/Switzerland/BE-ETHZ-260061/2020\|EPI_ISL_53 | ···················· | ···················· | ···················· | ···················· | ···················· | ···················· |
| H | 2153 | hCoV-19/Switzerland/ZH-ETHZ-260064/2020\|EPI_ISL_53 | ···················· | ···················· | ···················· | ···················· | ···················· | ···················· |
| H | 2154 | hCoV-19/Switzerland/VD-ETHZ-260069/2020\|EPI_ISL_53 | ···················· | ···················· | ···················· | ···················· | ···················· | ···················· |
| H | 2155 | hCoV-19/Switzerland/VD-ETHZ-260070/2020\|EPI_ISL_53 | ···················· | ···················· | ···················· | ···················· | ···················· | ···················· |
| H | 2156 | hCoV-19/Switzerland/BL-ETHZ-260075/2020\|EPI_ISL_53 | ···················· | ···················· | ···················· | ···················· | ···················· | ···················· |
| H | 2157 | hCoV-19/Switzerland/VD-ETHZ-260076/2020\|EPI_ISL_53 | ···················· | ···················· | ···················· | ···················· | ···················· | ···················· |
| H | 2158 | hCoV-19/Switzerland/VD-ETHZ-260077/2020\|EPI_ISL_53 | ···················· | ···················· | ···················· | ···················· | ···················· | ···················· |
| H | 2159 | hCoV-19/England/QEUH-96F640/2020\|EPI_ISL_540300\|20 | ···················· | ···················· | ···················· | ···················· | ···················· | ···················· |
| H | 2160 | hCoV-19/Scotland/QEUH-96FBD8/2020\|EPI_ISL_540301\|2 | ···················· | ···················· | ···················· | ···················· | ···················· | ···················· |
| H | 2161 | hCoV-19/Northern Ireland/QEUH-96FFBE/2020\|EPI_ISL_ | ···················· | ···················· | ···················· | ···················· | ···················· | ···················· |
| H | 2162 | hCoV-19/England/QEUH-96FA62/2020\|EPI_ISL_540303\|20 | ···················· | ···················· | ···················· | ···················· | ···················· | ···················· |
| H | 2163 | hCoV-19/England/QEUH-96F482/2020\|EPI_ISL_540304\|20 | ···················· | ···················· | ···················· | ···················· | ···················· | ···················· |
| H | 2164 | hCoV-19/England/QEUH-96F82C/2020\|EPI_ISL_540306\|20 | ···················· | ···················· | ···················· | ···················· | ···················· | ···················· |
| H | 2165 | hCoV-19/Northern Ireland/QEUH-96FEFD/2020\|EPI_ISL_ | ···················· | ···················· | ···················· | ···················· | ···················· | ···················· |
| H | 2166 | hCoV-19/England/QEUH-96F74D/2020\|EPI_ISL_540308\|20 | ···················· | ···················· | ···················· | ···················· | ···················· | ···················· |
| H | 2167 | hCoV-19/Northern Ireland/QEUH-97004E/2020\|EPI_ISL_ | ···················· | ···················· | ···················· | ···················· | ···················· | ···················· |
| H | 2168 | hCoV-19/England/QEUH-96FA80/2020\|EPI_ISL_540311\|20 | ···················· | ···················· | ···················· | ···················· | ···················· | ···················· |
| H | 2169 | hCoV-19/England/QEUH-96FA08/2020\|EPI_ISL_540312\|20 | ···················· | ···················· | ···················· | ···················· | ···················· | ···················· |
| H | 2170 | hCoV-19/England/QEUH-9700A8/2020\|EPI_ISL_540314\|20 | ···················· | ···················· | ···················· | ···················· | ···················· | ···················· |
| H | 2171 | hCoV-19/Scotland/QEUH-96F983/2020\|EPI_ISL_540315\|2 | ···················· | ···················· | ···················· | ···················· | ···················· | ···················· |
| H | 2172 | hCoV-19/England/QEUH-97008A/2020\|EPI_ISL_540316\|20 | ···················· | ···················· | ···················· | ···················· | ···················· | ···················· |
| H | 2173 | hCoV-19/Northern Ireland/QEUH-96FE2A/2020\|EPI_ISL_ | ···················· | ···················· | ···················· | ···················· | ···················· | ···················· |
| H | 2174 | hCoV-19/Scotland/QEUH-96FE0C/2020\|EPI_ISL_540318\|2 | ···················· | ···················· | ···················· | ···················· | ···················· | ···················· |
| H | 2175 | hCoV-19/England/QEUH-96FAEA/2020\|EPI_ISL_540319\|20 | ···················· | ···················· | ···················· | ···················· | ···················· | ···················· |
| H | 2176 | hCoV-19/England/QEUH-96F90B/2020\|EPI_ISL_540320\|20 | ···················· | ···················· | ···················· | ···················· | ···················· | ···················· |
| H | 2177 | hCoV-19/Scotland/QEUH-98F3AD/2020\|EPI_ISL_540366\|2 | ···················· | ···················· | ···················· | ···················· | ···················· | ···················· |
| H | 2178 | hCoV-19/Scotland/QEUH-98F7DE/2020\|EPI_ISL_540367\|2 | ···················· | ···················· | ···················· | ···················· | ···················· | ···················· |
| H | 2179 | hCoV-19/Scotland/QEUH-98F46E/2020\|EPI_ISL_540368\|2 | ···················· | ···················· | ···················· | ···················· | ···················· | ···················· |
| H | 2180 | hCoV-19/England/QEUH-98F2BF/2020\|EPI_ISL_540369\|20 | ···················· | ···················· | ···················· | ···················· | ···················· | ···················· |
| H | 2181 | hCoV-19/England/QEUH-98F194/2020\|EPI_ISL_540370\|20 | ···················· | ···················· | ···················· | ···················· | ···················· | ···················· |
| H | 2182 | hCoV-19/Scotland/QEUH-98EECB/2020\|EPI_ISL_540372\|2 | ···················· | ···················· | ···················· | ···················· | ···················· | ···················· |
| H | 2183 | hCoV-19/England/QEUH-98EFF5/2020\|EPI_ISL_540373\|20 | ···················· | ···················· | ···················· | ···················· | ···················· | ···················· |
| H | 2184 | hCoV-19/Scotland/QEUH-98F3E9/2020\|EPI_ISL_540376\|2 | ···················· | ···················· | ···················· | ···················· | ···················· | ···················· |
| H | 2185 | hCoV-19/Scotland/QEUH-98F149/2020\|EPI_ISL_540377\|2 | ···················· | ···················· | ···················· | ···················· | ···················· | ···················· |
| H | 2186 | hCoV-19/England/QEUH-98F4C8/2020\|EPI_ISL_540378\|20 | ···················· | ···················· | ···················· | ···················· | ···················· | ···················· |
| H | 2187 | hCoV-19/England/QEUH-98F282/2020\|EPI_ISL_540379\|20 | ···················· | ···················· | ···················· | ···················· | ···················· | ···················· |
| H | 2188 | hCoV-19/England/QEUH-98F2EC/2020\|EPI_ISL_540380\|20 | ···················· | ···················· | ···················· | ···················· | ···················· | ···················· |
| H | 2189 | hCoV-19/Scotland/QEUH-98F7CF/2020\|EPI_ISL_540381\|2 | ···················· | ···················· | ···················· | ···················· | ···················· | ···················· |
| H | 2190 | hCoV-19/Scotland/QEUH-98EEE9/2020\|EPI_ISL_540382\|2 | ···················· | ···················· | ···················· | ···················· | ···················· | ···················· |
| H | 2191 | hCoV-19/Scotland/QEUH-98F5E3/2020\|EPI_ISL_540383\|2 | ···················· | ···A················ | ···················· | ···················· | ···················· | ···················· |
| H | 2192 | hCoV-19/England/QEUH-98EE52/2020\|EPI_ISL_540384\|20 | ···················· | ···················· | ···················· | ···················· | ···················· | ···················· |
| H | 2193 | hCoV-19/England/QEUH-98EE43/2020\|EPI_ISL_540385\|20 | ···················· | ···················· | ···················· | ···················· | ···················· | ···················· |
| H | 2194 | hCoV-19/Scotland/QEUH-98F158/2020\|EPI_ISL_540386\|2 | ···················· | ···················· | ···················· | ···················· | ···················· | ···················· |

| | | | | | | | | |
|---|---|---|---|---|---|---|---|---|
| H | 2195 | hCoV-19/England/QEUH-98F4B9/2020\|EPI_ISL_540387\|20 | ···················· | ···················· | ···················· | ···················· | ···················· | ···················· |
| H | 2196 | hCoV-19/England/QEUH-98F228/2020\|EPI_ISL_540389\|20 | ···················· | ···················· | ···················· | ···················· | ···················· | ···················· |
| H | 2197 | hCoV-19/England/QEUH-98F316/2020\|EPI_ISL_540390\|20 | ···················· | ···················· | ···················· | ···················· | ···················· | ···················· |
| H | 2198 | hCoV-19/Scotland/QEUH-98F54D/2020\|EPI_ISL_540391\|2 | ···················· | ···················· | ···················· | ···················· | ···················· | ···················· |
| H | 2199 | hCoV-19/England/QEUH-98F097/2020\|EPI_ISL_540392\|20 | ···················· | ···················· | ···················· | ···················· | ···················· | ···················· |
| H | 2200 | hCoV-19/England/QEUH-98EEF8/2020\|EPI_ISL_540393\|20 | ···················· | ···················· | ···················· | ···················· | ···················· | ···················· |
| H | 2201 | hCoV-19/England/QEUH-98EE8F/2020\|EPI_ISL_540394\|20 | ···················· | ···················· | ···················· | ···················· | ···················· | ···················· |
| H | 2202 | hCoV-19/England/QEUH-98EF6E/2020\|EPI_ISL_540395\|20 | ···················· | ·A·················· | ···················· | ···················· | ···················· | ···················· |
| H | 2203 | hCoV-19/England/QEUH-98F307/2020\|EPI_ISL_540396\|20 | ···················· | ···················· | ···················· | ···················· | ···················· | ···················· |
| H | 2204 | hCoV-19/Scotland/QEUH-98F088/2020\|EPI_ISL_540397\|2 | ···················· | ···················· | ···················· | ···················· | ···················· | ···················· |
| H | 2205 | hCoV-19/England/QEUH-98EE9E/2020\|EPI_ISL_540398\|20 | ···················· | ···················· | ···················· | ···················· | ···················· | ···················· |
| H | 2206 | hCoV-19/England/QEUH-98EF13/2020\|EPI_ISL_540399\|20 | ···················· | ···················· | ···················· | ···················· | ···················· | ···················· |
| H | 2207 | hCoV-19/USA/WI-UW-1468/2020\|EPI_ISL_541600\|2020-09 | ···················· | ···················· | ···················· | ···················· | ···················· | ···················· |
| H | 2208 | hCoV-19/USA/WI-UW-1469/2020\|EPI_ISL_541601\|2020-09 | ···················· | ···················· | ···················· | ···················· | ···················· | ···················· |
| H | 2209 | hCoV-19/USA/WI-UW-1470/2020\|EPI_ISL_541602\|2020-09 | ···················· | ···················· | ···················· | ···················· | ···················· | ···················· |
| H | 2210 | hCoV-19/USA/WI-UW-1471/2020\|EPI_ISL_541603\|2020-09 | ···················· | ···················· | ···················· | ···················· | ···················· | ···················· |
| H | 2211 | hCoV-19/USA/WI-UW-1472/2020\|EPI_ISL_541604\|2020-09 | ···················· | ···················· | ···················· | ···················· | ···················· | ···················· |
| H | 2212 | hCoV-19/USA/WI-UW-1473/2020\|EPI_ISL_541605\|2020-09 | ···················· | ···················· | ···················· | ···················· | ···················· | ···················· |
| H | 2215 | hCoV-19/USA/WI-UW-1476/2020\|EPI_ISL_541608\|2020-09 | ···················· | ···················· | ···················· | ···················· | ···················· | ···················· |
| H | 2216 | hCoV-19/USA/WI-UW-1477/2020\|EPI_ISL_541609\|2020-09 | ···················· | ···················· | ···················· | ···················· | ···················· | ···················· |
| H | 2217 | hCoV-19/USA/WI-UW-1478/2020\|EPI_ISL_541610\|2020-09 | ···················· | ···················· | ···················· | ···················· | ···················· | ···················· |
| H | 2218 | hCoV-19/USA/WI-UW-1479/2020\|EPI_ISL_541611\|2020-09 | ···················· | ···················· | ···················· | ···················· | ···················· | ···················· |
| H | 2219 | hCoV-19/USA/WI-UW-1480/2020\|EPI_ISL_541612\|2020-09 | ···················· | ···················· | ···················· | ···················· | ···················· | ···················· |
| H | 2220 | hCoV-19/USA/WI-UW-1482/2020\|EPI_ISL_541613\|2020-09 | ···················· | ···················· | ···················· | ···················· | ···················· | ···················· |
| H | 2221 | hCoV-19/USA/WI-UW-1484/2020\|EPI_ISL_541614\|2020-09 | ···················· | ···················· | ···················· | ···················· | ···················· | ···················· |
| H | 2222 | hCoV-19/USA/WI-UW-1485/2020\|EPI_ISL_541615\|2020-09 | ···················· | ···················· | ···················· | ···················· | ···················· | ···················· |
| H | 2223 | hCoV-19/USA/WI-UW-1486/2020\|EPI_ISL_541616\|2020-09 | ···················· | ···················· | ···················· | ···················· | ···················· | ···················· |
| H | 2224 | hCoV-19/USA/WI-UW-1487/2020\|EPI_ISL_541617\|2020-09 | ···················· | ···················· | ···················· | ···················· | ···················· | ···················· |
| H | 2225 | hCoV-19/USA/WI-UW-1489/2020\|EPI_ISL_541618\|2020-09 | ···················· | ···················· | ···················· | ···················· | ···················· | ···················· |
| H | 2226 | hCoV-19/USA/WI-UW-1491/2020\|EPI_ISL_541619\|2020-09 | ···················· | ···················· | ···················· | ···················· | ···················· | ···················· |
| H | 2227 | hCoV-19/USA/WI-UW-1492/2020\|EPI_ISL_541620\|2020-09 | ···················· | ···················· | ···················· | ···················· | ···················· | ···················· |
| H | 2228 | hCoV-19/USA/WI-UW-1493/2020\|EPI_ISL_541621\|2020-09 | ···················· | ···················· | ···················· | ···················· | ···················· | ···················· |
| H | 2229 | hCoV-19/USA/WI-UW-1496/2020\|EPI_ISL_541623\|2020-09 | ···················· | ···················· | ···················· | ···················· | ···················· | ···················· |
| H | 2230 | hCoV-19/USA/WI-UW-1497/2020\|EPI_ISL_541624\|2020-09 | ···················· | ···················· | ···················· | ···················· | ···················· | ···················· |
| H | 2231 | hCoV-19/USA/WI-UW-1498/2020\|EPI_ISL_541625\|2020-09 | ···················· | ···················· | ···················· | ···················· | ···················· | ···················· |
| H | 2232 | hCoV-19/USA/WI-UW-1499/2020\|EPI_ISL_541626\|2020-09 | ···················· | ···················· | ···················· | ···················· | ···················· | ···················· |
| H | 2233 | hCoV-19/USA/WI-UW-1500/2020\|EPI_ISL_541627\|2020-09 | ···················· | ···················· | ···················· | ···················· | ···················· | ···················· |
| H | 2234 | hCoV-19/USA/WI-UW-1501/2020\|EPI_ISL_541628\|2020-09 | ···················· | ···················· | ···················· | ···················· | ···················· | ···················· |
| H | 2235 | hCoV-19/USA/WI-UW-1502/2020\|EPI_ISL_541629\|2020-09 | ···················· | ···················· | ···················· | ···················· | ···················· | ···················· |
| H | 2236 | hCoV-19/USA/WI-UW-1504/2020\|EPI_ISL_541630\|2020-09 | ···················· | ···················· | ···················· | ···················· | ···················· | ···················· |
| H | 2237 | hCoV-19/USA/WI-UW-1505/2020\|EPI_ISL_541631\|2020-09 | ···················· | ···················· | ···················· | ···················· | ···················· | ···················· |
| H | 2238 | hCoV-19/USA/WI-UW-1506/2020\|EPI_ISL_541632\|2020-09 | ···················· | ···················· | ···················· | ···················· | ···················· | ···················· |
| H | 2239 | hCoV-19/USA/WI-UW-1507/2020\|EPI_ISL_541633\|2020-09 | ···················· | ···················· | ···················· | ···················· | ···················· | ···················· |
| H | 2240 | hCoV-19/USA/WI-UW-1509/2020\|EPI_ISL_541635\|2020-09 | ···················· | ···················· | ···················· | ···················· | ···················· | ···················· |
| H | 2241 | hCoV-19/USA/WI-UW-1510/2020\|EPI_ISL_541636\|2020-09 | ···················· | ···················· | ···················· | ···················· | ···TT··············· | ···················· |
| H | 2242 | hCoV-19/USA/WI-UW-1511/2020\|EPI_ISL_541637\|2020-09 | ···················· | ···················· | ···················· | ···················· | ···················· | ···················· |
| H | 2243 | hCoV-19/USA/WI-UW-1512/2020\|EPI_ISL_541638\|2020-09 | ···················· | ···················· | ···················· | ···················· | ···················· | ···················· |
| H | 2244 | hCoV-19/USA/WI-UW-1513/2020\|EPI_ISL_541639\|2020-09 | ···················· | ···················· | ···················· | ···················· | ···················· | ···················· |
| H | 2246 | hCoV-19/USA/WI-UW-1515/2020\|EPI_ISL_541641\|2020-09 | ···················· | ···················· | ···················· | ···················· | ···················· | ···················· |
| H | 2248 | hCoV-19/USA/WI-UW-1517/2020\|EPI_ISL_541643\|2020-09 | ···················· | ·A·················· | ···················· | ···················· | ···················· | ···················· |
| H | 2249 | hCoV-19/USA/WI-UW-1519/2020\|EPI_ISL_541644\|2020-09 | ···················· | ···················· | ···················· | ···················· | ···················· | ···················· |
| H | 2251 | hCoV-19/USA/WI-UW-1523/2020\|EPI_ISL_541646\|2020-09 | ···················· | ···················· | ···················· | ···················· | ···················· | ···················· |
| H | 2252 | hCoV-19/USA/WI-UW-1525/2020\|EPI_ISL_541647\|2020-09 | ···················· | ···················· | ···················· | ···················· | ···················· | ···················· |
| H | 2253 | hCoV-19/USA/WI-UW-1526/2020\|EPI_ISL_541648\|2020-09 | ···················· | ···················· | ···················· | ···················· | ···················· | ···················· |
| H | 2254 | hCoV-19/Serbia/NP-0440907/2020\|EPI_ISL_541649\|2020 | ···················· | ···················· | ···················· | ···················· | ···················· | ···················· |
| H | 2255 | hCoV-19/Serbia/NP-0401307/2020\|EPI_ISL_541650\|2020 | ···················· | ···················· | ···················· | ···················· | ···················· | ···················· |
| H | 2256 | hCoV-19/Serbia/KM-0370707/2020\|EPI_ISL_541651\|2020 | ···················· | ···················· | ···················· | ···················· | ···················· | ···················· |
| H | 2257 | hCoV-19/Serbia/NP-011507/2020\|EPI_ISL_541652\|2020- | ···················· | ···················· | ···················· | ···················· | ···················· | ···················· |
| H | 2258 | hCoV-19/India/MH-NIV-13768/2020\|EPI_ISL_541697\|202 | ···················· | ···················· | ···················· | ···················· | ···················· | ···················· |
| H | 2260 | hCoV-19/India/MH-NIV-138100/2020\|EPI_ISL_541699\|20 | ···················· | ···················· | ···················· | ···················· | ···················· | ···················· |
| H | 2261 | hCoV-19/USA/TX-HMH-1916/2020\|EPI_ISL_542900\|2020-0 | ···················· | ···················· | ···················· | ···················· | ···················· | ···················· |
| H | 2262 | hCoV-19/USA/TX-HMH-1917/2020\|EPI_ISL_542901\|2020-0 | ···················· | ···················· | ···················· | ···················· | ···················· | ···················· |
| H | 2263 | hCoV-19/USA/TX-HMH-1919/2020\|EPI_ISL_542902\|2020-0 | ···················· | ···················· | ···················· | ···················· | ···················· | ···················· |
| H | 2264 | hCoV-19/USA/TX-HMH-1924/2020\|EPI_ISL_542904\|2020-0 | ···················· | ···················· | ···················· | ···················· | ···················· | ···················· |
| H | 2265 | hCoV-19/USA/TX-HMH-1926/2020\|EPI_ISL_542905\|2020-0 | ···················· | ···················· | ···················· | ···················· | ···················· | ···················· |
| H | 2266 | hCoV-19/USA/TX-HMH-1928/2020\|EPI_ISL_542906\|2020-0 | ···················· | ···················· | ···················· | ···················· | ···················· | ···················· |
| H | 2267 | hCoV-19/USA/TX-HMH-1931/2020\|EPI_ISL_542907\|2020-0 | ···················· | ···················· | ···················· | ···················· | ···················· | ···················· |
| H | 2268 | hCoV-19/USA/TX-HMH-1935/2020\|EPI_ISL_542908\|2020-0 | ···················· | ···················· | ···················· | ···················· | ···················· | ···················· |
| H | 2269 | hCoV-19/USA/TX-HMH-1936/2020\|EPI_ISL_542909\|2020-0 | ···················· | ···················· | ···················· | ···················· | ···················· | ···················· |
| H | 2270 | hCoV-19/USA/TX-HMH-1938/2020\|EPI_ISL_542910\|2020-0 | ···················· | ···················· | ···················· | ···················· | ···················· | ···················· |
| H | 2271 | hCoV-19/USA/TX-HMH-1939/2020\|EPI_ISL_542911\|2020-0 | ···················· | ···················· | ···················· | ···················· | ···················· | ···················· |
| H | 2272 | hCoV-19/USA/TX-HMH-1945/2020\|EPI_ISL_542913\|2020-0 | ···················· | ···················· | ···················· | ···················· | ···················· | ···················· |
| H | 2273 | hCoV-19/USA/TX-HMH-1947/2020\|EPI_ISL_542914\|2020-0 | ···················· | ···················· | ···················· | ···················· | ···················· | ···················· |
| H | 2274 | hCoV-19/USA/TX-HMH-1949/2020\|EPI_ISL_542915\|2020-0 | ···················· | ···················· | ···················· | ···················· | ···················· | ···················· |
| H | 2275 | hCoV-19/USA/TX-HMH-1950/2020\|EPI_ISL_542916\|2020-0 | ···················· | ···················· | ···················· | ···················· | ···················· | ···················· |
| H | 2276 | hCoV-19/USA/TX-HMH-1951/2020\|EPI_ISL_542917\|2020-0 | ···················· | ··············C····· | ···················· | ···················· | ···················· | ···················· |
| H | 2277 | hCoV-19/USA/TX-HMH-1952/2020\|EPI_ISL_542918\|2020-0 | ···················· | ··············C····· | ···················· | ···················· | ···················· | ···················· |
| H | 2278 | hCoV-19/USA/TX-HMH-1956/2020\|EPI_ISL_542920\|2020-0 | ···················· | ···················· | ···················· | ···················· | ···················· | ···················· |
| H | 2279 | hCoV-19/USA/TX-HMH-1960/2020\|EPI_ISL_542921\|2020-0 | ···················· | ······A············· | ···················· | ···················· | ···················· | ···················· |
| H | 2280 | hCoV-19/USA/TX-HMH-1961/2020\|EPI_ISL_542922\|2020-0 | ···················· | ···················· | ···················· | ···················· | ···················· | ···················· |

| | | | | | | | | | | |
|---|---|---|---|---|---|---|---|---|---|---|
| H | 2281 | hCoV-19/USA/TX-HMH-1962/2020|EPI_ISL_542923|2020-0 | ···················· | ······A············· | ···················· | ···················· | ···················· | ···················· | | |
| H | 2282 | hCoV-19/USA/TX-HMH-1964/2020|EPI_ISL_542924|2020-0 | ···················· | ···················· | ···················· | ···················· | ···················· | ···················· | | |
| H | 2283 | hCoV-19/USA/TX-HMH-1968/2020|EPI_ISL_542925|2020-0 | ···················· | ···················· | ···················· | ···················· | ···················· | ···················· | | |
| H | 2284 | hCoV-19/USA/TX-HMH-1969/2020|EPI_ISL_542926|2020-0 | ···················· | ···················· | ···················· | ···················· | ···················· | ···················· | | |
| H | 2285 | hCoV-19/USA/TX-HMH-1970/2020|EPI_ISL_542927|2020-0 | ···················· | ···················· | ···················· | ···················· | ···················· | ···················· | | |
| H | 2286 | hCoV-19/USA/TX-HMH-1973/2020|EPI_ISL_542929|2020-0 | ···················· | ···················· | ···················· | ···················· | ···················· | ···············A···· | | |
| H | 2287 | hCoV-19/USA/TX-HMH-1974/2020|EPI_ISL_542930|2020-0 | ···················· | ···················· | ···················· | ···················· | ···················· | ···················· | | |
| H | 2288 | hCoV-19/USA/TX-HMH-1975/2020|EPI_ISL_542931|2020-0 | ···················· | ···················· | ···················· | ···················· | ···················· | ···················· | | |
| H | 2289 | hCoV-19/USA/TX-HMH-1981/2020|EPI_ISL_542934|2020-0 | ···················· | ···················· | ···················· | ···················· | ···················· | ···················· | | |
| H | 2290 | hCoV-19/USA/NM-UNM-00529/2020|EPI_ISL_542951|2020- | ···················· | ···················· | ···················· | ···················· | ···················· | ···················· | | |
| H | 2291 | hCoV-19/USA/TX-HMH-3918/2020|EPI_ISL_543280|2020-0 | ···················· | ···················· | ···················· | ···················· | ···················· | ···················· | | |
| H | 2292 | hCoV-19/USA/TX-HMH-3921/2020|EPI_ISL_543281|2020-0 | ···················· | ···················· | ···················· | ···················· | ···················· | ···················· | | |
| H | 2293 | hCoV-19/USA/TX-HMH-3923/2020|EPI_ISL_543282|2020-0 | ···················· | ···················· | ···················· | ···················· | ···················· | ···················· | | |
| H | 2294 | hCoV-19/USA/TX-HMH-3924/2020|EPI_ISL_543283|2020-0 | ···················· | ···················· | ···················· | ···················· | ···················· | ···················· | | |
| H | 2295 | hCoV-19/USA/TX-HMH-3926/2020|EPI_ISL_543285|2020-0 | ···················· | ···················· | ···················· | ···················· | ···················· | ···················· | | |
| H | 2300 | hCoV-19/USA/TX-HMH-4009/2020|EPI_ISL_543292|2020-0 | ···················· | ···················· | ···················· | ···················· | ···················· | ···················· | | |
| H | 2301 | hCoV-19/USA/TX-HMH-4010/2020|EPI_ISL_543293|2020-0 | ···················· | ···················· | ···················· | ···················· | ···················· | ··N··N··········N | 1 | 1 |
| H | 2302 | hCoV-19/USA/TX-HMH-4011/2020|EPI_ISL_543294|2020-0 | ···················· | ···················· | ···················· | ···················· | ···················· | ···················· | | |
| H | 2303 | hCoV-19/USA/TX-HMH-4016/2020|EPI_ISL_543298|2020-0 | ···················· | ···················· | ···················· | ···················· | ···················· | ···················· | | |
| H | 2305 | hCoV-19/Scotland/QEUH-964D82/2020|EPI_ISL_531214|2 | ···················· | ···················· | ···················· | ···················· | ···················· | ···················· | | |
| H | 2306 | hCoV-19/England/QEUH-9695EF/2020|EPI_ISL_531215|20 | ···················· | ···················· | ···················· | ···················· | ···················· | ···················· | | |
| H | 2307 | hCoV-19/England/QEUH-969AA4/2020|EPI_ISL_531216|20 | ···················· | ···················· | ···················· | ···················· | ···················· | ···················· | | |
| H | 2308 | hCoV-19/England/QEUH-965817/2020|EPI_ISL_531217|20 | ···················· | ···················· | ···················· | ···················· | ···················· | ···················· | | |
| H | 2309 | hCoV-19/England/QEUH-969655/2020|EPI_ISL_531218|20 | ···················· | ···················· | ···················· | ···················· | ···················· | ···················· | | |
| H | 2310 | hCoV-19/Wales/QEUH-964933/2020|EPI_ISL_531219|2020 | ···················· | ···················· | ···················· | ···················· | ···················· | ···················· | | |
| H | 2311 | hCoV-19/Northern Ireland/QEUH-9698AA/2020|EPI_ISL_ | ···················· | ···················· | ···················· | ···················· | ···················· | ···················· | | |
| H | 2312 | hCoV-19/England/QEUH-964FE6/2020|EPI_ISL_531222|20 | ···················· | ···················· | ···················· | ···················· | ···················· | ···················· | | |
| H | 2313 | hCoV-19/Scotland/QEUH-964A30/2020|EPI_ISL_531223|2 | ···················· | ···················· | ···················· | ···················· | ···················· | ···················· | | |
| H | 2314 | hCoV-19/England/QEUH-96ADAA/2020|EPI_ISL_531226|20 | ···················· | ···················· | ···················· | ···················· | ···················· | ···················· | | |
| H | 2315 | hCoV-19/England/QEUH-96BCAC/2020|EPI_ISL_531227|20 | ···················· | ···················· | ···················· | ···················· | ···················· | ···················· | | |
| H | 2316 | hCoV-19/England/QEUH-96B143/2020|EPI_ISL_531229|20 | ···················· | ···················· | ···················· | ···················· | ···················· | ···················· | | |
| H | 2317 | hCoV-19/Northern Ireland/QEUH-96ABFB/2020|EPI_ISL_ | ···················· | ···················· | ···················· | ···················· | ···················· | ···················· | | |
| H | 2318 | hCoV-19/England/QEUH-96BDE5/2020|EPI_ISL_531232|20 | ···················· | ···················· | ···················· | ···················· | ···················· | ···················· | | |
| H | 2319 | hCoV-19/Northern Ireland/QEUH-96B87B/2020|EPI_ISL_ | ···················· | ···················· | ···················· | ···················· | ···················· | ···················· | | |
| H | 2320 | hCoV-19/Northern Ireland/QEUH-96AA2B/2020|EPI_ISL_ | ···················· | ···················· | ···················· | ···················· | ···················· | ···················· | | |
| H | 2321 | hCoV-19/England/QEUH-96B2C8/2020|EPI_ISL_531236|20 | ···················· | ···················· | ···················· | ···················· | ···················· | ···················· | | |
| H | 2322 | hCoV-19/Northern Ireland/QEUH-96B134/2020|EPI_ISL_ | ···················· | ···················· | ···················· | ···················· | ···················· | ···················· | | |
| H | 2323 | hCoV-19/Northern Ireland/QEUH-96AAFE/2020|EPI_ISL_ | ···················· | ···················· | ···················· | ···················· | ···················· | ···················· | | |
| H | 2324 | hCoV-19/England/QEUH-96AE98/2020|EPI_ISL_531240|20 | ···················· | ···················· | ···················· | ···················· | ···················· | ···················· | | |
| H | 2325 | hCoV-19/England/QEUH-96B671/2020|EPI_ISL_531241|20 | ···················· | ···················· | ···················· | ···················· | ···················· | ···················· | | |
| H | 2326 | hCoV-19/England/QEUH-96A988/2020|EPI_ISL_531242|20 | ···················· | ···················· | ···················· | ···················· | ···················· | ···················· | | |
| H | 2327 | hCoV-19/England/QEUH-96BB09/2020|EPI_ISL_531244|20 | ···················· | ···················· | ···················· | ···················· | ···················· | ···················· | | |
| H | 2328 | hCoV-19/England/QEUH-96B4A4/2020|EPI_ISL_531245|20 | ···················· | ···················· | ···················· | ···················· | ···················· | ···················· | | |
| H | 2329 | hCoV-19/England/QEUH-96BE79/2020|EPI_ISL_531247|20 | ···················· | ···················· | ···················· | ···················· | ···················· | ···················· | | |
| H | 2330 | hCoV-19/Scotland/QEUH-96B69F/2020|EPI_ISL_531249|2 | ···················· | ···················· | ···················· | ···················· | ···················· | ···················· | | |
| H | 2331 | hCoV-19/Wales/QEUH-96BE00/2020|EPI_ISL_531250|2020 | ···················· | ···················· | ···················· | ···················· | ···················· | ···················· | | |
| H | 2332 | hCoV-19/Northern Ireland/QEUH-96B85D/2020|EPI_ISL_ | ···················· | ···················· | ···················· | ···················· | ···················· | ···················· | | |
| H | 2333 | hCoV-19/England/QEUH-96B18F/2020|EPI_ISL_531253|20 | ···················· | ···················· | ···················· | ···················· | ···················· | ···················· | | |
| H | 2334 | hCoV-19/England/QEUH-96BCF7/2020|EPI_ISL_531254|20 | ···················· | ···················· | ···················· | ···················· | ···················· | ···················· | | |
| H | 2335 | hCoV-19/England/QEUH-96AD22/2020|EPI_ISL_531255|20 | ···················· | ···················· | ···················· | ···················· | ···················· | ···················· | | |
| H | 2336 | hCoV-19/Northern Ireland/QEUH-96AB55/2020|EPI_ISL_ | ···················· | ···················· | ···················· | ···················· | ···················· | ···················· | | |
| H | 2337 | hCoV-19/England/QEUH-96B7C9/2020|EPI_ISL_531257|20 | ···················· | ·A·················· | ···················· | ···················· | ···················· | ···················· | | |
| H | 2338 | hCoV-19/England/QEUH-96AEE3/2020|EPI_ISL_531258|20 | ···················· | ···················· | ···················· | ···················· | ···················· | ···················· | | |
| H | 2339 | hCoV-19/England/QEUH-96B583/2020|EPI_ISL_531259|20 | ···················· | ···················· | ···················· | ···················· | ···················· | ···················· | | |
| H | 2340 | hCoV-19/England/QEUH-96BE97/2020|EPI_ISL_531260|20 | ···················· | ···················· | ···················· | ···················· | ···················· | ···················· | | |
| H | 2341 | hCoV-19/Northern Ireland/QEUH-96B84E/2020|EPI_ISL_ | ···················· | ···················· | ···················· | ···················· | ···················· | ···················· | | |
| H | 2342 | hCoV-19/England/QEUH-96BDB8/2020|EPI_ISL_531262|20 | ···················· | ···················· | ···················· | ···················· | ···················· | ···················· | | |
| H | 2343 | hCoV-19/England/QEUH-96AE4D/2020|EPI_ISL_531263|20 | ···················· | ·A·················· | ···················· | ···················· | ···················· | ···················· | | |
| H | 2344 | hCoV-19/Northern Ireland/QEUH-96AB46/2020|EPI_ISL_ | ···················· | ···················· | ···················· | ···················· | ···················· | ···················· | | |
| H | 2345 | hCoV-19/Northern Ireland/QEUH-96B91E/2020|EPI_ISL_ | ···················· | ···················· | ···················· | ···················· | ···················· | ···················· | | |
| H | 2346 | hCoV-19/Scotland/QEUH-96B389/2020|EPI_ISL_531266|2 | ···················· | ···················· | ···················· | ···················· | ···················· | ···················· | | |
| H | 2347 | hCoV-19/Wales/QEUH-96B152/2020|EPI_ISL_531267|2020 | ···················· | ···················· | ···················· | ···················· | ···················· | ···················· | | |
| H | 2348 | hCoV-19/England/QEUH-96AF68/2020|EPI_ISL_531268|20 | ···················· | ···················· | ···················· | ···················· | ···················· | ···················· | | |
| H | 2349 | hCoV-19/England/QEUH-96BA57/2020|EPI_ISL_531269|20 | ···················· | ···················· | ···················· | ···················· | ···················· | ···················· | | |
| H | 2350 | hCoV-19/England/QEUH-96BD5E/2020|EPI_ISL_531270|20 | ···················· | ···················· | ···················· | ···················· | ···················· | ···················· | | |
| H | 2351 | hCoV-19/England/QEUH-96B996/2020|EPI_ISL_531271|20 | ···················· | ···················· | ···················· | ···················· | ···················· | ···················· | | |
| H | 2352 | hCoV-19/England/QEUH-96B714/2020|EPI_ISL_531272|20 | ···················· | ···················· | ···················· | ···················· | ···················· | ···················· | | |
| H | 2353 | hCoV-19/England/QEUH-96AEA7/2020|EPI_ISL_531273|20 | ···················· | ·A·················· | ···················· | ···················· | ···················· | ···················· | | |
| H | 2354 | hCoV-19/England/QEUH-96AED4/2020|EPI_ISL_531274|20 | ···················· | ···················· | ···················· | ···················· | ···················· | ···················· | | |
| H | 2355 | hCoV-19/England/QEUH-96BA48/2020|EPI_ISL_531275|20 | ···················· | ···················· | ···················· | ···················· | ···················· | ···················· | | |
| H | 2356 | hCoV-19/England/QEUH-96B41D/2020|EPI_ISL_531276|20 | ···················· | ···················· | ···················· | ···················· | ···················· | ···················· | | |
| H | 2357 | hCoV-19/England/QEUH-96AEC5/2020|EPI_ISL_531277|20 | ···················· | ·A·················· | ···················· | ···················· | ···················· | ···················· | | |
| H | 2358 | hCoV-19/England/QEUH-96B899/2020|EPI_ISL_531278|20 | ···················· | ···················· | ···················· | ···················· | ···················· | ···················· | | |
| H | 2359 | hCoV-19/England/QEUH-96BC7F/2020|EPI_ISL_531280|20 | ···················· | ···················· | ···················· | ···················· | ···················· | ···················· | | |
| H | 2360 | hCoV-19/England/QEUH-96B556/2020|EPI_ISL_531281|20 | ···················· | ···················· | ···················· | ···················· | ···················· | ···················· | | |
| H | 2361 | hCoV-19/England/QEUH-96B2F5/2020|EPI_ISL_531282|20 | ···················· | ···················· | ···················· | ···················· | ···················· | ···················· | | |
| H | 2362 | hCoV-19/Wales/QEUH-96BE2E/2020|EPI_ISL_531283|2020 | ···················· | ···················· | ···················· | ···················· | ···················· | ···················· | | |
| H | 2363 | hCoV-19/England/QEUH-96BBEB/2020|EPI_ISL_531284|20 | ···················· | ···················· | ···················· | ···················· | ···················· | ···················· | | |
| H | 2364 | hCoV-19/England/QEUH-96AEF2/2020|EPI_ISL_531285|20 | ···················· | ···················· | ···················· | ···················· | ···················· | ···················· | | |
| H | 2365 | hCoV-19/Northern Ireland/QEUH-96AA94/2020|EPI_ISL_ | ···················· | ···················· | ···················· | ···················· | ···················· | ···················· | | |

H 2366 hCoV-19/Northern Ireland/QEUH-96B8F3/2020|EPI_ISL_ ···················· ···················· ···················· ···················· ···················· ····················
H 2367 hCoV-19/England/QEUH-96B626/2020|EPI_ISL_531288|20 ···················· ·A·················· ···················· ···················· ···················· ····················
H 2368 hCoV-19/England/QEUH-96B32F/2020|EPI_ISL_531290|20 ···················· ···················· ···················· ···················· ···················· ····················
H 2369 hCoV-19/England/QEUH-96A979/2020|EPI_ISL_531292|20 ···················· ···················· ···················· ···················· ···················· ····················
H 2370 hCoV-19/England/QEUH-96BCD9/2020|EPI_ISL_531293|20 ···················· ···················· ···················· ···················· ···················· ····················
H 2371 hCoV-19/England/QEUH-96BED3/2020|EPI_ISL_531295|20 ···················· ···················· ···················· ···················· ···················· ····················
H 2372 hCoV-19/England/QEUH-96BD21/2020|EPI_ISL_531296|20 ···················· ···················· ···················· ···················· ···················· ····················
H 2373 hCoV-19/England/QEUH-96BFD0/2020|EPI_ISL_531297|20 ···················· ·A·················· ···················· ···················· ···················· ····················
H 2374 hCoV-19/England/QEUH-96AE3E/2020|EPI_ISL_531298|20 ···················· ·A·················· ···················· ···················· ···················· ····················
H 2375 hCoV-19/England/QEUH-96BAB1/2020|EPI_ISL_531299|20 ···················· ···················· ···················· ···················· ···················· ····················
H 2376 hCoV-19/England/QEUH-96BB81/2020|EPI_ISL_531300|20 ···················· ···················· ···················· ···················· ··················C· ···················· 1
H 2377 hCoV-19/Northern Ireland/QEUH-96AB19/2020|EPI_ISL_ ···················· ···················· ···················· ···················· ···················· ····················
H 2378 hCoV-19/Northern Ireland/QEUH-96AA1C/2020|EPI_ISL_ ···················· ···················· ···················· ···················· ···················· ····················
H 2379 hCoV-19/England/QEUH-943EB3/2020|EPI_ISL_532545|20 ···················· ···················· ···················· ···················· ······T············· ····················
H 2380 hCoV-19/England/QEUH-9435CD/2020|EPI_ISL_532548|20 ···················· ···················· ···················· ···················· ···················· ····················
H 2381 hCoV-19/England/QEUH-94367F/2020|EPI_ISL_532550|20 ···················· ···················· ···················· ···················· ···················· ····················
H 2382 hCoV-19/England/QEUH-943E2C/2020|EPI_ISL_532553|20 ···················· ···················· ···················· ···················· ···················· ····················
H 2383 hCoV-19/England/QEUH-94393A/2020|EPI_ISL_532554|20 ···················· ···················· ···················· ···················· ···················· ····················
H 2384 hCoV-19/Northern Ireland/QEUH-943DA7/2020|EPI_ISL_ ···················· ···················· ···················· ···················· ···················· ····················
H 2385 hCoV-19/England/QEUH-943EE0/2020|EPI_ISL_532556|20 ···················· ···················· ···················· ···················· ···················· ····················
H 2386 hCoV-19/England/QEUH-943C31/2020|EPI_ISL_532557|20 ···················· ···················· ···················· ···················· ···················· ····················
H 2387 hCoV-19/Northern Ireland/QEUH-943C40/2020|EPI_ISL_ ···················· ···················· ···················· ···················· ···················· ····················
H 2388 hCoV-19/England/QEUH-9434DF/2020|EPI_ISL_532559|20 ···················· ···················· ···················· ···················· ···················· ····················
H 2389 hCoV-19/Northern Ireland/QEUH-943493/2020|EPI_ISL_ ···················· ···················· ···················· ···················· ···················· ····················
H 2390 hCoV-19/Northern Ireland/QEUH-94385B/2020|EPI_ISL_ ···················· ···················· ···················· ···················· ···················· ····················
H 2391 hCoV-19/Scotland/QEUH-9439A3/2020|EPI_ISL_532563|2 ···················· ···················· ···················· ···················· ···················· ····················
H 2392 hCoV-19/Scotland/QEUH-94375E/2020|EPI_ISL_532564|2 ···················· ···················· ···················· ···················· ···················· ····················
H 2393 hCoV-19/England/QEUH-943ADD/2020|EPI_ISL_532565|20 ···················· ···················· ···················· ···················· ···················· ····················
H 2394 hCoV-19/Northern Ireland/QEUH-943BE9/2020|EPI_ISL_ ···················· ···················· ···················· ···················· ···················· ····················
H 2395 hCoV-19/Northern Ireland/QEUH-9434C0/2020|EPI_ISL_ ···················· ···················· ···················· ···················· ···················· ····················
H 2396 hCoV-19/England/QEUH-943A19/2020|EPI_ISL_532570|20 ···················· ···················· ···················· ···················· ···················· ····················
H 2397 hCoV-19/England/QEUH-943606/2020|EPI_ISL_532571|20 ···················· ···················· ···················· ···················· ···················· ····················
H 2398 hCoV-19/Scotland/QEUH-9436E8/2020|EPI_ISL_532572|2 ···················· ···················· ···················· ···················· ···A················ ····················
H 2399 hCoV-19/England/QEUH-9439FE/2020|EPI_ISL_532573|20 ···················· ···················· ···················· ···················· ···················· ····················
H 2400 hCoV-19/England/QEUH-943387/2020|EPI_ISL_532574|20 ···················· ···················· ···················· ···················· ···················· ····················
H 2401 hCoV-19/Northern Ireland/QEUH-943DC5/2020|EPI_ISL_ ···················· ···················· ···················· ···················· ···················· ····················
H 2402 hCoV-19/England/QEUH-9433E1/2020|EPI_ISL_532576|20 ···················· ···················· ···················· ···················· ···················· ····················
H 2403 hCoV-19/England/QEUH-943E68/2020|EPI_ISL_532577|20 ···················· ···················· ···················· ···················· ···················· ····················
H 2404 hCoV-19/Northern Ireland/QEUH-94390D/2020|EPI_ISL_ ···················· ···················· ···················· ···················· ···················· ····················
H 2405 hCoV-19/Northern Ireland/QEUH-9438F1/2020|EPI_ISL_ ···················· ···················· ···················· ···················· ···················· ····················
H 2406 hCoV-19/England/QEUH-943B25/2020|EPI_ISL_532581|20 ···················· ···················· ···················· ···················· ···················· ····················
H 2407 hCoV-19/Scotland/QEUH-94384C/2020|EPI_ISL_532582|2 ···················· ···················· ···················· ···················· ···················· ····················
H 2408 hCoV-19/England/QEUH-943633/2020|EPI_ISL_532584|20 ···················· ···················· ···················· ···················· ···················· ····················
H 2409 hCoV-19/England/QEUH-943AEC/2020|EPI_ISL_532585|20 ···················· ···················· ···················· ···················· ···················· ····················
H 2410 hCoV-19/England/QEUH-943ED1/2020|EPI_ISL_532586|20 ···················· ···················· ···················· ···················· ···················· ····················
H 2411 hCoV-19/Scotland/QEUH-9435AF/2020|EPI_ISL_532587|2 ···················· ···················· ···················· ···················· ···················· ····················
H 2412 hCoV-19/Northern Ireland/QEUH-9434FD/2020|EPI_ISL_ ···················· ···················· ···················· ···················· ···················· ····················
H 2413 hCoV-19/England/QEUH-9437E5/2020|EPI_ISL_532589|20 ···················· ···················· ···················· ···················· ···················· ····················
H 2414 hCoV-19/England/QEUH-9433D2/2020|EPI_ISL_532590|20 ···················· ···················· ···················· ···················· ···················· ····················
H 2415 hCoV-19/England/QEUH-943518/2020|EPI_ISL_532591|20 ···················· ···················· ···················· ···················· ···················· ····················
H 2416 hCoV-19/England/QEUH-943E59/2020|EPI_ISL_532592|20 ···················· ···················· ···················· ···················· ···················· ····················
H 2417 hCoV-19/England/QEUH-943509/2020|EPI_ISL_532593|20 ···················· ···················· ···················· ···················· ···················· ····················
H 2418 hCoV-19/Scotland/QEUH-9439C1/2020|EPI_ISL_532594|2 ···················· ···················· ···················· ···················· ···················· ····················
H 2419 hCoV-19/Northern Ireland/QEUH-94386A/2020|EPI_ISL_ ···················· ···················· ···················· ···················· ···················· ····················
H 2420 hCoV-19/England/QEUH-943642/2020|EPI_ISL_532597|20 ···················· ···················· ···················· ···················· ···················· ····················
H 2421 hCoV-19/Scotland/QEUH-943712/2020|EPI_ISL_532598|2 ···················· ···················· ···················· ···················· ···A················ ····················
H 2422 hCoV-19/England/QEUH-943545/2020|EPI_ISL_532599|20 ···················· ···················· ···················· ···················· ···················· ····················
H 2423 hCoV-19/Northern Ireland/QEUH-9434A2/2020|EPI_ISL_ ···················· ···················· ···················· ···················· ···················· ····················
H 2424 hCoV-19/England/QEUH-9435FA/2020|EPI_ISL_532601|20 ···················· ···················· ···················· ···················· ···················· ····················
H 2425 hCoV-19/England/QEUH-94340C/2020|EPI_ISL_532602|20 ···················· ···················· ···················· ···················· ···················· ····················
H 2426 hCoV-19/England/QEUH-943949/2020|EPI_ISL_532603|20 ···················· ···················· ···················· ···················· ···················· ····················
H 2427 hCoV-19/England/QEUH-94342A/2020|EPI_ISL_532604|20 ···················· ···················· ···················· ···················· ···················· ····················
H 2428 hCoV-19/England/QEUH-943651/2020|EPI_ISL_532605|20 ···················· ···················· ···················· ···················· ···················· ····················
H 2429 hCoV-19/England/QEUH-94378B/2020|EPI_ISL_532607|20 ···················· ···················· ···················· ···················· ···················· ····················
H 2430 hCoV-19/England/QEUH-94335A/2020|EPI_ISL_532608|20 ···················· ···················· ···················· ···················· ···················· ····················
H 2431 hCoV-19/England/QEUH-943976/2020|EPI_ISL_532609|20 ···················· ···················· ···················· ···················· ···················· ····················
H 2432 hCoV-19/England/QEUH-943581/2020|EPI_ISL_532610|20 ···················· ···················· ···················· ···················· ···················· ····················
H 2433 hCoV-19/England/QEUH-94379A/2020|EPI_ISL_532613|20 ···················· ···················· ···················· ···················· ···················· ····················
H 2434 hCoV-19/Northern Ireland/QEUH-943D98/2020|EPI_ISL_ ···················· ···················· ···················· ···················· ···················· ····················
H 2435 hCoV-19/Northern Ireland/QEUH-9434EE/2020|EPI_ISL_ ···················· ···················· ···················· ···················· ···················· ····················
H 2436 hCoV-19/England/QEUH-9436D9/2020|EPI_ISL_532616|20 ···················· ···················· ···················· ···················· ···················· ····················
H 2437 hCoV-19/England/QEUH-943B70/2020|EPI_ISL_532617|20 ···················· ···················· ···················· ···················· ···················· ····················
H 2438 hCoV-19/England/QEUH-943615/2020|EPI_ISL_532618|20 ···················· ···················· ···················· ···················· ···················· ····················
H 2439 hCoV-19/England/QEUH-9435EB/2020|EPI_ISL_532619|20 ···················· ···················· ···················· ···················· ···················· ····················
H 2440 hCoV-19/Scotland/QEUH-943703/2020|EPI_ISL_532620|2 ···················· ···················· ···················· ···················· ···A················ ····················
H 2441 hCoV-19/Scotland/QEUH-943439/2020|EPI_ISL_532621|2 ···················· ···················· ···················· ···················· ···················· ····················
H 2442 hCoV-19/England/QEUH-94377C/2020|EPI_ISL_532622|20 ···················· ···················· ···················· ···················· ···················· ····················
H 2443 hCoV-19/England/QEUH-9433F0/2020|EPI_ISL_532623|20 ···················· ···················· ···················· ···················· ···················· ····················
H 2444 hCoV-19/Northern Ireland/QEUH-9438E2/2020|EPI_ISL_ ···················· ···················· ···················· ···················· ···················· ····················
H 2445 hCoV-19/Scotland/QEUH-9437B8/2020|EPI_ISL_532625|2 ···················· ···················· ···················· ···················· ···················· ····················

H 2446 hCoV-19/Scotland/QEUH-943ABF/2020|EPI_ISL_532626|2 ···················· ···················· ···················· ···················· ···················· ····················
H 2447 hCoV-19/England/QEUH-943C6E/2020|EPI_ISL_532627|20 ···················· ···················· ···················· ···················· ···················· ····················
H 2448 hCoV-19/England/QEUH-943E1D/2020|EPI_ISL_532628|20 ···················· ···················· ···················· ···················· ···················· ····················
H 2449 hCoV-19/England/QEUH-94368E/2020|EPI_ISL_532629|20 ···················· ···················· ···················· ···················· ···················· ····················
H 2450 hCoV-19/England/QEUH-943484/2020|EPI_ISL_532630|20 ···················· ···················· ··········T········· ···················· ···················· ····················
H 2451 hCoV-19/Wales/QEUH-9411AD/2020|EPI_ISL_532631|2020 ···················· ···················· ···················· ···················· ···················· ····················
H 2452 hCoV-19/England/QEUH-941125/2020|EPI_ISL_532632|20 ···················· ···················· ···················· ···················· ···················· ····················
H 2453 hCoV-19/Northern Ireland/QEUH-9410BF/2020|EPI_ISL_ ···················· ···················· ···················· ···················· ···················· ····················
H 2454 hCoV-19/Canada/QC-LSPQ-L00235561/2020|EPI_ISL_5361 ···················· ···················· ···················· ···················· ···················· ····················
H 2455 hCoV-19/Canada/QC-LSPQ-L00235607/2020|EPI_ISL_5361 ···················· ···················· ···················· ···················· ···················· ····················
H 2456 hCoV-19/Canada/QC-LSPQ-L00235639/2020|EPI_ISL_5361 ···················· ···················· ···················· ···················· ···················· ····················
H 2457 hCoV-19/Canada/QC-LSPQ-L00235671/2020|EPI_ISL_5361 ···················· ···················· ···················· ···················· ···················· ····················
H 2458 hCoV-19/Canada/QC-LSPQ-L00235772/2020|EPI_ISL_5361 ···················· ···················· ···················· ···················· ···················· ····················
H 2459 hCoV-19/Canada/QC-LSPQ-L00235800/2020|EPI_ISL_5361 ···················· ···················· ···················· ···················· ···················· ····················
H 2460 hCoV-19/Canada/QC-LSPQ-L00237841/2020|EPI_ISL_5361 ···················· ···················· ···················· ···················· ···················· ····················
H 2461 hCoV-19/Canada/QC-LSPQ-L00237878/2020|EPI_ISL_5361 ···················· ···················· ···················· ···················· ···················· ····················
H 2462 hCoV-19/Canada/QC-LSPQ-L00237886/2020|EPI_ISL_5361 ···················· ···················· ···················· ···················· ···················· ····················
H 2463 hCoV-19/Canada/QC-LSPQ-L00237888/2020|EPI_ISL_5361 ···················· ···················· ···················· ···················· ···················· ····················
H 2466 hCoV-19/Spain/RI-IBV-99005884/2020|EPI_ISL_537426| ···················· ···················· ···················· ···················· ···················· ····················
H 2470 hCoV-19/Spain/RI-IBV-99005889/2020|EPI_ISL_537431| ···················· ···················· ···················· ···················· ···················· ····················
H 2472 hCoV-19/Spain/RI-IBV-99005892/2020|EPI_ISL_537434| ···················· ···················· ···················· ···················· ···················· ····················
H 2473 hCoV-19/Spain/RI-IBV-99005894/2020|EPI_ISL_537436| ···················· ···················· ···················· ···················· ·NN················· ····················
H 2474 hCoV-19/Spain/RI-IBV-99005897/2020|EPI_ISL_537437| ···················· ···················· ···················· ···················· ···················· ····················
H 2476 hCoV-19/Spain/RI-IBV-99005900/2020|EPI_ISL_537439| ···················· ···················· ···················· ···················· ···················· ····················
H 2477 hCoV-19/England/LEED-2A96F6/2020|EPI_ISL_538750|20 ···················· ·A·················· ···················· ···················· ···················· ····················
H 2478 hCoV-19/England/LEED-2A973F/2020|EPI_ISL_538751|20 ···················· ·A·················· ···················· ···················· ···················· ····················
H 2479 hCoV-19/England/LEED-2A974E/2020|EPI_ISL_538752|20 ···················· ·A·················· ···················· ···················· ···················· ····················
H 2480 hCoV-19/England/LEED-2A967E/2020|EPI_ISL_538753|20 ···················· ···················A ···················· ···················· ···················· ···················· 1
H 2481 hCoV-19/England/LEED-2A981E/2020|EPI_ISL_538755|20 ···················· ···················· ···················· ···················· ···················· ····················
H 2482 hCoV-19/England/LEED-2AA015/2020|EPI_ISL_538756|20 ···················· ···················· ···················· ···················· ···················· ····················
H 2483 hCoV-19/England/LEED-2A9799/2020|EPI_ISL_538757|20 ···················· ···················· ···················· ···················· ···················· ····················
H 2484 hCoV-19/England/LEED-2A9FCE/2020|EPI_ISL_538758|20 ···················· ···················· ···················· ···················· ···················· ····················
H 2485 hCoV-19/England/LEED-2A9F82/2020|EPI_ISL_538759|20 ···················· ···················· ···················· ···················· ···················· ····················
H 2486 hCoV-19/England/LEED-2A97F3/2020|EPI_ISL_538760|20 ···················· ···················· ···················· ···················· ···················· ····················
H 2487 hCoV-19/England/LEED-2A9F73/2020|EPI_ISL_538761|20 ···················· ·A·················· ···················· ···················· ···················· ····················
H 2488 hCoV-19/England/LEED-2A9EC1/2020|EPI_ISL_538762|20 ···················· ···················· ···················· ···················· ···················· ····················
H 2489 hCoV-19/England/LEED-2A9711/2020|EPI_ISL_538764|20 ···················· ·A·················· ···················· ···················· ···················· ····················
H 2490 hCoV-19/England/LEED-2A9EA3/2020|EPI_ISL_538765|20 ···················· ···················· ···················· ···················· ···················· ····················
H 2491 hCoV-19/England/LEED-2A9E94/2020|EPI_ISL_538766|20 ···················· ·A·················· ···················· ···················· ···················· ····················
H 2492 hCoV-19/England/LEED-2A9FFB/2020|EPI_ISL_538767|20 ···················· ·A·················· ···················· ···················· ···················· ····················
H 2493 hCoV-19/England/LEED-2A9F91/2020|EPI_ISL_538768|20 ···················· ·A·················· ···················· ···················· ···················· ····················
H 2494 hCoV-19/England/LEED-2A9DD3/2020|EPI_ISL_538769|20 ···················· ···················· ···················· ···················· ···················· ····················
H 2495 hCoV-19/USA/TX-HMH-3796/2020|EPI_ISL_543200|2020-0 ···················· ···················· ···················· ···················· ···················· ····················
H 2501 hCoV-19/USA/TX-HMH-3803/2020|EPI_ISL_543207|2020-0 ···················· ···················· ···················· ···················· ···················· ····················
H 2502 hCoV-19/USA/TX-HMH-3804/2020|EPI_ISL_543208|2020-0 ···················· ·············A······ ···················· ···················· ···················· ····················
H 2508 hCoV-19/USA/TX-HMH-3810/2020|EPI_ISL_543214|2020-0 ···················· ···················· ···················· ···················· ···················· ····················
H 2511 hCoV-19/USA/TX-HMH-3813/2020|EPI_ISL_543217|2020-0 ···················· ···················· ···················· ···················· ···················· ····················
H 2513 hCoV-19/USA/TX-HMH-3815/2020|EPI_ISL_543219|2020-0 ···················· ···················· ···················· ···················· ···················· ····················
H 2515 hCoV-19/USA/TX-HMH-3817/2020|EPI_ISL_543221|2020-0 ···················· ···················· ···················· ···················· ··············N····· NN··················
H 2517 hCoV-19/USA/TX-HMH-3819/2020|EPI_ISL_543223|2020-0 ···················· ·············A······ ···················· ···················· ···················· ····················
H 2521 hCoV-19/USA/TX-HMH-3823/2020|EPI_ISL_543227|2020-0 ···················· ···················· ···················· ···················· ···················· ····················
H 2523 hCoV-19/USA/TX-HMH-3827/2020|EPI_ISL_543231|2020-0 ···················· ···················· ···················· ···················· ···················· ····················
H 2525 hCoV-19/USA/TX-HMH-3830/2020|EPI_ISL_543233|2020-0 ···················· ···················· ···················· ···················· ···················· ····················
H 2527 hCoV-19/USA/TX-HMH-3832/2020|EPI_ISL_543235|2020-0 ···················· ···················· ···················· ···················· ···················· ····················
H 2528 hCoV-19/USA/TX-HMH-3834/2020|EPI_ISL_543237|2020-0 ···················· ···················· ···················· ···················· ···················· ····················
H 2529 hCoV-19/USA/TX-HMH-3836/2020|EPI_ISL_543239|2020-0 ···················· ···················· ···················· ···················· ···················· ····················
H 2531 hCoV-19/USA/TX-HMH-3838/2020|EPI_ISL_543241|2020-0 ···················· ···················· ···················· ···················· ···················· ····················
H 2536 hCoV-19/USA/TX-HMH-3843/2020|EPI_ISL_543246|2020-0 ···················· ···················· ···················· ···················· ···················· ····················
H 2540 hCoV-19/USA/TX-HMH-3850/2020|EPI_ISL_543252|2020-0 ···················· ···················· ···················· ···················· ···················· ····················
H 2541 hCoV-19/USA/TX-HMH-3851/2020|EPI_ISL_543253|2020-0 ···················· ···················· ···················· ···················· ···················· ····················
H 2544 hCoV-19/USA/TX-HMH-3855/2020|EPI_ISL_543256|2020-0 ···················· ···················· ···················· ···················· ···················· ····················
H 2545 hCoV-19/USA/TX-HMH-3858/2020|EPI_ISL_543259|2020-0 ···················· ···················· ···················· ···················· ···················· ····················
H 2546 hCoV-19/USA/TX-HMH-3859/2020|EPI_ISL_543260|2020-0 ···················· ···················· ···················· ···················· ···················· ····················
H 2547 hCoV-19/USA/TX-HMH-3861/2020|EPI_ISL_543261|2020-0 ···················· ···················· ···················· ···················· ···················· ····················
H 2548 hCoV-19/USA/TX-HMH-3864/2020|EPI_ISL_543263|2020-0 ···················· ···················· ···················· ···················· ···················· ····················
H 2549 hCoV-19/USA/TX-HMH-3865/2020|EPI_ISL_543264|2020-0 ···················· ···················· ···················· ···················· ···················· ····················
H 2550 hCoV-19/USA/TX-HMH-3869/2020|EPI_ISL_543266|2020-0 ···················· ···················· ···················· ···················· ···················· ····················
H 2551 hCoV-19/USA/TX-HMH-3870/2020|EPI_ISL_543267|2020-0 ···················· ···················· ···················· ···················· ···················· ····················
H 2552 hCoV-19/USA/TX-HMH-3876/2020|EPI_ISL_543269|2020-0 ···················· ···················· ···················· ···················· ···················· ····················
H 2553 hCoV-19/USA/TX-HMH-3877/2020|EPI_ISL_543270|2020-0 ········T··········· ···················· ···················· ···················· ···················· ····················
H 2554 hCoV-19/USA/TX-HMH-3878/2020|EPI_ISL_543271|2020-0 ···················· ·············A······ ···················· ···················· ···················· ····················
H 2555 hCoV-19/USA/TX-HMH-3903/2020|EPI_ISL_543272|2020-0 ···················· ···················· ···················· ···················· ···················· ····················
H 2557 hCoV-19/USA/TX-HMH-3910/2020|EPI_ISL_543275|2020-0 ···················· ·············A······ ···················· ···················· ···················· ····················
H 2558 hCoV-19/USA/TX-HMH-3913/2020|EPI_ISL_543276|2020-0 ···················· ···················· ···················· ···················· ···················· ····················
H 2559 hCoV-19/USA/TX-HMH-3914/2020|EPI_ISL_543277|2020-0 ···················· ···················· ···················· ···················· ···················· ····················
H 2560 hCoV-19/USA/TX-HMH-3915/2020|EPI_ISL_543278|2020-0 ···················· ···················· ···················· ···················· ···················· ····················
H 2561 hCoV-19/England/QEUH-9652FB/2020|EPI_ISL_531180|20 ···················· ···················· ···················· ···················· ···················· ····················
H 2562 hCoV-19/England/QEUH-96538F/2020|EPI_ISL_531181|20 ···················· ···················· ···················· ···················· ···················· ····················
H 2563 hCoV-19/England/QEUH-9696BF/2020|EPI_ISL_531183|20 ···················· ···················· ···················· ···················· ···················· ····················
H 2564 hCoV-19/Northern Ireland/QEUH-964F04/2020|EPI_ISL_ ···················· ···················· ···················· ···················· ···················· ····················

| | | | | | | | |
|---|---|---|---|---|---|---|---|
| H | 2565 hCoV-19/England/QEUH-969B38/2020\|EPI_ISL_531185\|20 | ···················· | ·A·················· | ···················· | ···················· | ···················· | ···················· |
| H | 2566 hCoV-19/England/QEUH-9694A6/2020\|EPI_ISL_531186\|20 | ···················· | ·A·················· | ···················· | ···················· | ···················· | ···················· |
| H | 2567 hCoV-19/Scotland/QEUH-969743/2020\|EPI_ISL_531189\|2 | ···················· | ···················· | ···················· | ···················· | ···················· | ···················· |
| H | 2568 hCoV-19/England/QEUH-9656B3/2020\|EPI_ISL_531192\|20 | ···················· | ···················· | ···················· | ···················· | ···················· | ···················· |
| H | 2569 hCoV-19/Northern Ireland/QEUH-96501F/2020\|EPI_ISL_ | ···················· | ···················· | ···················· | ···················· | ···················· | ···················· |
| H | 2570 hCoV-19/Northern Ireland/QEUH-964E52/2020\|EPI_ISL_ | ···················· | ···················· | ···················· | ···················· | ···················· | ···················· |
| H | 2571 hCoV-19/England/QEUH-9649E8/2020\|EPI_ISL_531195\|20 | ···················· | ·A·················· | ···················· | ···················· | ···················· | ··A················· |
| H | 2572 hCoV-19/England/QEUH-965000/2020\|EPI_ISL_531196\|20 | ···················· | ···················· | ···················· | ···················· | ···················· | ···················· |
| H | 2573 hCoV-19/Northern Ireland/QEUH-964E8F/2020\|EPI_ISL_ | ···················· | ···················· | ···················· | ···················· | ···················· | ···················· |
| H | 2574 hCoV-19/Scotland/QEUH-965738/2020\|EPI_ISL_531198\|2 | ···················· | ···················· | ···················· | ···················· | ···················· | ···················· |
| H | 2575 hCoV-19/England/QEUH-9657DE/2020\|EPI_ISL_531199\|20 | ···················· | ···················· | ···················· | ···················· | ···················· | ···················· |
| H | 2576 hCoV-19/Northern Ireland/QEUH-96AA49/2020\|EPI_ISL_ | ···················· | ···················· | ···················· | ···················· | ···················· | ···················· |
| H | 2577 hCoV-19/England/QEUH-96B29B/2020\|EPI_ISL_531305\|20 | ···················· | ···················· | ···················· | ···················· | ···················· | ···················· |
| H | 2578 hCoV-19/Northern Ireland/QEUH-96B86C/2020\|EPI_ISL_ | ···················· | ···················· | ···················· | ···················· | ···················· | ···················· |
| H | 2579 hCoV-19/Northern Ireland/QEUH-96AC25/2020\|EPI_ISL_ | ···················· | ···················· | ···················· | ···················· | ···················· | ···················· |
| H | 2580 hCoV-19/England/QEUH-96AFB3/2020\|EPI_ISL_531308\|20 | ···················· | ···················· | ···················· | ···················· | ···················· | ···················· |
| H | 2581 hCoV-19/England/QEUH-96BD30/2020\|EPI_ISL_531309\|20 | ···················· | ·A·················· | ···················· | ···················· | ···················· | ··A················· |
| H | 2582 hCoV-19/England/QEUH-96B082/2020\|EPI_ISL_531310\|20 | ···················· | ···················· | ···················· | ···················· | ···················· | ···················· |
| H | 2583 hCoV-19/England/QEUH-96BA66/2020\|EPI_ISL_531311\|20 | ···················· | ···················· | ···················· | ···················· | ···················· | ···················· |
| H | 2584 hCoV-19/England/QEUH-96B3D4/2020\|EPI_ISL_531312\|20 | ···················· | ···················· | ···················· | ···················· | ···················· | ···················· |
| H | 2585 hCoV-19/England/QEUH-96BEB5/2020\|EPI_ISL_531313\|20 | ···················· | ···················· | ···················· | ···················· | ···················· | ···················· |
| H | 2586 hCoV-19/Northern Ireland/QEUH-96AB37/2020\|EPI_ISL_ | ···················· | ···················· | ···················· | ···················· | ···················· | ···················· |
| H | 2587 hCoV-19/Northern Ireland/QEUH-96B969/2020\|EPI_ISL_ | ···················· | ···················· | ···················· | ···················· | ···················· | ···················· |
| H | 2588 hCoV-19/England/QEUH-96AA3A/2020\|EPI_ISL_531316\|20 | ···················· | ···················· | ···················· | ···················· | ···················· | ···················· |
| H | 2589 hCoV-19/Northern Ireland/QEUH-96AC61/2020\|EPI_ISL_ | ···················· | ···················· | ···················· | ···················· | ···················· | ···················· |
| H | 2590 hCoV-19/England/QEUH-96B107/2020\|EPI_ISL_531318\|20 | ···················· | ···················· | ···················· | ···················· | ···················· | ···················· |
| H | 2591 hCoV-19/England/QEUH-96BBDC/2020\|EPI_ISL_531319\|20 | ···················· | ···················· | ···················· | ···················· | ···················· | ···················· |
| H | 2592 hCoV-19/Northern Ireland/QEUH-96AB0A/2020\|EPI_ISL_ | ···················· | ···················· | ···················· | ···················· | ···················· | ···················· |
| H | 2593 hCoV-19/Scotland/QEUH-96B1BC/2020\|EPI_ISL_531321\|2 | ···················· | ···················· | ···················· | ···················· | ···················· | ···················· |
| H | 2594 hCoV-19/England/QEUH-96AF4A/2020\|EPI_ISL_531322\|20 | ···················· | ···················· | ···················· | ···················· | ···················· | ···················· |
| H | 2595 hCoV-19/England/QEUH-96ABA0/2020\|EPI_ISL_531323\|20 | ···················· | ···················· | ···················· | ···················· | ·····T·············· | ···················· |
| H | 2596 hCoV-19/Northern Ireland/QEUH-96AF86/2020\|EPI_ISL_ | ···················· | ···················· | ···················· | ···················· | ···················· | ···················· |
| H | 2597 hCoV-19/England/QEUH-96ADB9/2020\|EPI_ISL_531325\|20 | ···················· | ···················· | ···················· | ···················· | ···················· | ···················· |
| H | 2598 hCoV-19/England/QEUH-96AD8C/2020\|EPI_ISL_531326\|20 | ···················· | ···················· | ···················· | ···················· | ···················· | ···················· |
| H | 2599 hCoV-19/England/QEUH-96BD4F/2020\|EPI_ISL_531327\|20 | ···················· | ···················· | ···················· | ···················· | ···················· | ···················· |
| H | 2600 hCoV-19/England/QEUH-96B4D1/2020\|EPI_ISL_531329\|20 | ···················· | ···················· | ···················· | ···················· | ···················· | ···················· |
| H | 2601 hCoV-19/Scotland/QEUH-96AE5C/2020\|EPI_ISL_531330\|2 | ···················· | ···················· | ···················· | ···················· | ···················· | ···················· |
| H | 2602 hCoV-19/Northern Ireland/QEUH-96B90F/2020\|EPI_ISL_ | ···················· | ···················· | ···················· | ···················· | ···················· | ···················· |
| H | 2603 hCoV-19/England/QEUH-96AFC2/2020\|EPI_ISL_531332\|20 | ···················· | ···················· | ···················· | ···················· | ···················· | ···················· |
| H | 2604 hCoV-19/England/QEUH-96B8C6/2020\|EPI_ISL_531334\|20 | ···················· | ·A·················· | ···················· | ···················· | ···················· | ···················· |
| H | 2605 hCoV-19/England/QEUH-94127D/2020\|EPI_ISL_532635\|20 | ···················· | ···················· | ···················· | ···················· | ···················· | ···················· |
| H | 2606 hCoV-19/England/QEUH-941170/2020\|EPI_ISL_532637\|20 | ···················· | ···················· | ···················· | ···················· | ···················· | ···················· |
| H | 2607 hCoV-19/England/QEUH-94119E/2020\|EPI_ISL_532641\|20 | ···················· | ···················· | ···················· | ···················· | ···················· | ···················· |
| H | 2608 hCoV-19/England/QEUH-94128C/2020\|EPI_ISL_532642\|20 | ···················· | ···················· | ···················· | ···················· | ···················· | ···················· |
| H | 2609 hCoV-19/Northern Ireland/QEUH-941213/2020\|EPI_ISL_ | ···················· | ···················· | ···················· | ···················· | ···················· | ···················· |
| H | 2610 hCoV-19/England/QEUH-94126E/2020\|EPI_ISL_532644\|20 | ···················· | ···················· | ···················· | ···················· | ···················· | ···················· |
| H | 2611 hCoV-19/Wales/QEUH-941161/2020\|EPI_ISL_532647\|2020 | ···················· | ···················· | ···················· | ···················· | ···················· | ···················· |
| H | 2612 hCoV-19/Scotland/QEUH-9410FB/2020\|EPI_ISL_532648\|2 | ···················· | ···················· | ···················· | ···················· | ···················· | ···················· |
| H | 2613 hCoV-19/Wales/QEUH-94129B/2020\|EPI_ISL_532649\|2020 | ···················· | ···················· | ···················· | ···················· | ···················· | ···················· |
| H | 2614 hCoV-19/England/QEUH-8835E0/2020\|EPI_ISL_532650\|20 | ···················· | ···················· | ···················· | ···················· | ···················· | ···················· |
| H | 2615 hCoV-19/England/QEUH-8AEDC9/2020\|EPI_ISL_532651\|20 | ···················· | ···················· | ···················· | ···················· | ···················· | ···················· |
| H | 2616 hCoV-19/England/QEUH-89A847/2020\|EPI_ISL_532652\|20 | ···················· | ···················· | ···················· | ···················· | ···················· | ···················· |
| H | 2617 hCoV-19/England/QEUH-8845D0/2020\|EPI_ISL_532654\|20 | ···················· | ···················· | ···················· | ···················· | ···················· | ···················· |
| H | 2618 hCoV-19/Northern Ireland/QEUH-88F159/2020\|EPI_ISL_ | ···················· | ···················· | ···················· | ···················· | ···················· | ···················· |
| H | 2619 hCoV-19/England/QEUH-882059/2020\|EPI_ISL_532657\|20 | ···················· | ···················· | ···················· | ···················· | ···················· | ···················· |
| H | 2620 hCoV-19/Scotland/QEUH-89D1C1/2020\|EPI_ISL_532659\|2 | ···················· | ···················· | ···················· | ···················· | ···················· | ···················· |
| H | 2621 hCoV-19/England/QEUH-880149/2020\|EPI_ISL_532660\|20 | ···················· | ···················· | ···················· | ···················· | ···················· | ···················· |
| H | 2622 hCoV-19/Northern Ireland/QEUH-8A42F1/2020\|EPI_ISL_ | ···················· | ···················· | ···················· | ···················· | ···················· | ···················· |
| H | 2623 hCoV-19/England/QEUH-89E9AA/2020\|EPI_ISL_532663\|20 | ···················· | ···················· | ···················· | ···················· | ···················· | ···················· |
| H | 2624 hCoV-19/England/QEUH-8AFBFB/2020\|EPI_ISL_532665\|20 | ···················· | ···················· | ···················· | ···················· | ···················· | ···················· |
| H | 2625 hCoV-19/Canada/QC-LSPQ-L00235802/2020\|EPI_ISL_5361 | ···················· | ···················· | ···················· | ···················· | ···················· | ···················· |
| H | 2626 hCoV-19/Canada/QC-LSPQ-L00235815/2020\|EPI_ISL_5361 | ···················· | ···················· | ···················· | ···················· | ···················· | ···················· |
| H | 2627 hCoV-19/Canada/QC-LSPQ-L00235849/2020\|EPI_ISL_5361 | ···················· | ···················· | ···················· | ···················· | ···················· | ···················· |
| H | 2628 hCoV-19/Canada/QC-LSPQ-L00235855/2020\|EPI_ISL_5361 | ···················· | ···················· | ···················· | ···················· | ···················· | ···················· |
| H | 2629 hCoV-19/Canada/QC-LSPQ-L00235866/2020\|EPI_ISL_5361 | ···················· | ···················· | ···················· | ···················· | ···················· | ···················· |
| H | 2630 hCoV-19/Canada/QC-LSPQ-L00235907/2020\|EPI_ISL_5361 | ···················· | ···················· | ···················· | ···················· | ···················· | ···················· |
| H | 2631 hCoV-19/Canada/QC-LSPQ-L00235965/2020\|EPI_ISL_5361 | ···················· | ···················· | ···················· | ···················· | ···················· | ···················· |
| H | 2632 hCoV-19/Canada/QC-LSPQ-L00236091/2020\|EPI_ISL_5361 | ···················· | ···················· | ···················· | ···················· | ···················· | ···················· |
| H | 2633 hCoV-19/Canada/QC-LSPQ-L00236212/2020\|EPI_ISL_5361 | ···················· | ···················· | ···················· | ···················· | ···················· | ···················· |
| H | 2634 hCoV-19/Canada/QC-LSPQ-L00236215/2020\|EPI_ISL_5361 | ···················· | ···················· | ···················· | ···················· | ···················· | ···················· |
| H | 2635 hCoV-19/Canada/QC-LSPQ-L00236258/2020\|EPI_ISL_5361 | ···················· | ···················· | ···················· | ···················· | ···················· | ···················· |
| H | 2636 hCoV-19/Canada/QC-LSPQ-L00236367/2020\|EPI_ISL_5361 | ···················· | ···················· | ···················· | ···················· | ···················· | ···················· |
| H | 2637 hCoV-19/Canada/QC-LSPQ-L00236394/2020\|EPI_ISL_5361 | ···················· | ···················· | ···················· | ···················· | ···················· | ···················· |
| H | 2638 hCoV-19/Canada/QC-LSPQ-L00236396/2020\|EPI_ISL_5361 | ···················· | ···················· | ···················· | ···················· | ···················· | ···················· |
| H | 2639 hCoV-19/Canada/QC-LSPQ-L00236403/2020\|EPI_ISL_5361 | ···················· | ···················· | ···················· | ···················· | ···················· | ···················· |
| H | 2640 hCoV-19/Canada/QC-LSPQ-L00236426/2020\|EPI_ISL_5361 | ···················· | ···················· | ···················· | ···················· | ···················· | ···················· |
| H | 2641 hCoV-19/Canada/QC-LSPQ-L00236430/2020\|EPI_ISL_5361 | ···················· | ···················· | ···················· | ···················· | ···················· | ···················· |
| H | 2642 hCoV-19/Canada/QC-LSPQ-L00236439/2020\|EPI_ISL_5361 | ···················· | ···················· | ···················· | ···················· | ···················· | ···················· |
| H | 2643 hCoV-19/Canada/QC-LSPQ-L00236526/2020\|EPI_ISL_5361 | ···················· | ·A·················· | ···················· | ···················· | ···················· | ···················· |
| H | 2644 hCoV-19/Canada/QC-LSPQ-L00236536/2020\|EPI_ISL_5361 | ···················· | ···················· | ···················· | ···················· | ···················· | ···················· |

| | | | | | | | |
|---|---|---|---|---|---|---|---|
| H | 2645 hCoV-19/Canada/QC-LSPQ-L00236558/2020\|EPI_ISL_5361 | ···················· | ···················· | ···················· | ···················· | ···················· | ···················· |
| H | 2646 hCoV-19/Canada/QC-LSPQ-L00236573/2020\|EPI_ISL_5361 | ···················· | ···················· | ···················· | ···················· | ···················· | ···················· |
| H | 2647 hCoV-19/Canada/QC-LSPQ-L00236582/2020\|EPI_ISL_5361 | ···················· | ···················· | ···················· | ···················· | ···················· | ···················· |
| H | 2648 hCoV-19/Canada/QC-LSPQ-L00236595/2020\|EPI_ISL_5361 | ···················· | ···················· | ···················· | ···················· | ···················· | ···················· |
| H | 2649 hCoV-19/Canada/QC-LSPQ-L00236598/2020\|EPI_ISL_5361 | ···················· | ···················· | ···················· | ···················· | ···················· | ···················· |
| H | 2650 hCoV-19/Canada/QC-LSPQ-L00236601/2020\|EPI_ISL_5361 | ···················· | ···················· | ···················· | ···················· | ···················· | ···················· |
| H | 2651 hCoV-19/Canada/QC-LSPQ-L00236620/2020\|EPI_ISL_5361 | ···················· | ···················· | ···················· | ···················· | ···················· | ···················· |
| H | 2652 hCoV-19/Canada/QC-LSPQ-L00236653/2020\|EPI_ISL_5361 | ···················· | ···················· | ···················· | ···················· | ···················· | ···················· |
| H | 2653 hCoV-19/Canada/QC-LSPQ-L00236657/2020\|EPI_ISL_5361 | ···················· | ···················· | ···················· | ···················· | ···················· | ···················· |
| H | 2654 hCoV-19/Canada/QC-LSPQ-L00236779/2020\|EPI_ISL_5361 | ···················· | ···················· | ···················· | ···················· | ···················· | ···················· |
| H | 2655 hCoV-19/Canada/QC-LSPQ-L00236781/2020\|EPI_ISL_5361 | ···················· | ···················· | ···················· | ···················· | ···················· | ···················· |
| H | 2656 hCoV-19/Canada/QC-LSPQ-L00236805/2020\|EPI_ISL_5361 | ···················· | ···················· | ···················· | ···················· | ···················· | ···················· |
| H | 2657 hCoV-19/Canada/QC-LSPQ-L00236905/2020\|EPI_ISL_5361 | ···················· | ···················· | ···················· | ···················· | ···················· | ···················· |
| H | 2658 hCoV-19/Canada/QC-LSPQ-L00236922/2020\|EPI_ISL_5361 | ···················· | ···················· | ···················· | ···················· | ···················· | ···················· |
| H | 2659 hCoV-19/Canada/QC-LSPQ-L00236940/2020\|EPI_ISL_5361 | ···················· | ···················· | ···················· | ···················· | ···················· | ···················· |
| H | 2660 hCoV-19/Canada/QC-LSPQ-L00236943/2020\|EPI_ISL_5361 | ···················· | ···················· | ···················· | ···················· | ···················· | ···················· |
| H | 2661 hCoV-19/Canada/QC-LSPQ-L00236973/2020\|EPI_ISL_5361 | ···················· | ···················· | ···················· | ···················· | ···················· | ···················· |
| H | 2662 hCoV-19/Canada/QC-LSPQ-L00237011/2020\|EPI_ISL_5361 | ···················· | ···················· | ···················· | ···················· | ···················· | ···················· |
| H | 2663 hCoV-19/Canada/QC-LSPQ-L00237078/2020\|EPI_ISL_5361 | ···················· | ···················· | ···················· | ···················· | ···················· | ···················· |
| H | 2664 hCoV-19/Canada/QC-LSPQ-L00237080/2020\|EPI_ISL_5361 | ···················· | ···················· | ···················· | ···················· | ···················· | ···················· |
| H | 2665 hCoV-19/Canada/QC-LSPQ-L00237091/2020\|EPI_ISL_5361 | ···················· | ···················· | ···················· | ···················· | ···················· | ···················· |
| H | 2666 hCoV-19/Canada/QC-LSPQ-L00237108/2020\|EPI_ISL_5361 | ···················· | ···················· | ···················· | ···················· | ···················· | ···················· |
| H | 2667 hCoV-19/Canada/QC-LSPQ-L00237109/2020\|EPI_ISL_5361 | ···················· | ···················· | ···················· | ···················· | ···················· | ···················· |
| H | 2668 hCoV-19/Canada/QC-LSPQ-L00237135/2020\|EPI_ISL_5361 | ···················· | ···················· | ···················· | ···················· | ···················· | ···················· |
| H | 2669 hCoV-19/Canada/QC-LSPQ-L00237217/2020\|EPI_ISL_5361 | ···················· | ·A·················· | ···················· | ···················· | ···················· | ···················· |
| H | 2670 hCoV-19/Canada/QC-LSPQ-L00237242/2020\|EPI_ISL_5361 | ···················· | ·A·················· | ···················· | ···················· | ···················· | ···················· |
| H | 2671 hCoV-19/Canada/QC-LSPQ-L00237287/2020\|EPI_ISL_5361 | ···················· | ···················· | ···················· | ···················· | ···················· | ···················· |
| H | 2672 hCoV-19/Canada/QC-LSPQ-L00237312/2020\|EPI_ISL_5361 | ···················· | ···················· | ···················· | ···················· | ···················· | ···················· |
| H | 2673 hCoV-19/Canada/QC-LSPQ-L00237340/2020\|EPI_ISL_5361 | ···················· | ···················· | ···················· | ···················· | ···················· | ···················· |
| H | 2674 hCoV-19/Canada/QC-LSPQ-L00237365/2020\|EPI_ISL_5361 | ···················· | ···················· | ···················· | ···················· | ···················· | ···················· |
| H | 2675 hCoV-19/Canada/QC-LSPQ-L00237372/2020\|EPI_ISL_5361 | ···················· | ···················· | ···················· | ···················· | ···················· | ···················· |
| H | 2676 hCoV-19/Canada/QC-LSPQ-L00237374/2020\|EPI_ISL_5361 | ···················· | ···················· | ···················· | ···················· | ···················· | ···················· |
| H | 2677 hCoV-19/Canada/QC-LSPQ-L00237419/2020\|EPI_ISL_5361 | ···················· | ···················· | ···················· | ···················· | ···················· | ···················· |
| H | 2678 hCoV-19/Canada/QC-LSPQ-L00237465/2020\|EPI_ISL_5361 | ···················· | ···················· | ···················· | ···················· | ···················· | ···················· |
| H | 2679 hCoV-19/Canada/QC-LSPQ-L00237490/2020\|EPI_ISL_5361 | ···················· | ···················· | ···················· | ···················· | ···················· | ···················· |
| H | 2680 hCoV-19/Canada/QC-LSPQ-L00237503/2020\|EPI_ISL_5361 | ···················· | ···················· | ···················· | ···················· | ···················· | ···················· |
| H | 2681 hCoV-19/Canada/QC-LSPQ-L00237577/2020\|EPI_ISL_5361 | ···················· | ···················· | ···················· | ···················· | ···················· | ···················· |
| H | 2682 hCoV-19/Canada/QC-LSPQ-L00237589/2020\|EPI_ISL_5361 | ···················· | ···················· | ···················· | ···················· | ···················· | ···················· |
| H | 2683 hCoV-19/Spain/RI-IBV-99005901/2020\|EPI_ISL_537440\| | ···················· | ···················· | ···················· | ···················· | ···················· | ···················· |
| H | 2684 hCoV-19/Spain/RI-IBV-99005905/2020\|EPI_ISL_537443\| | ···················· | ···················· | ···················· | ···················· | ··N················· | ···················· |
| H | 2687 hCoV-19/Spain/RI-IBV-99005909/2020\|EPI_ISL_537447\| | ···················· | ···················· | ···················· | ···················· | ···················· | ···················· |
| H | 2690 hCoV-19/Spain/RI-IBV-99005915/2020\|EPI_ISL_537453\| | ···················· | ···················· | ···················· | ···················· | ···················· | ···················· |
| H | 2694 hCoV-19/USA/CA-UCLA-1/2020\|EPI_ISL_537467\|2020-04- | ···················· | ···················· | ···················· | ···················· | ···················· | ···················· |
| H | 2695 hCoV-19/USA/CA-UCLA-105/2020\|EPI_ISL_537469\|2020-0 | ···················· | ···················· | ···················· | ···················· | ···················· | ···················· |
| H | 2696 hCoV-19/USA/CA-UCLA-11/2020\|EPI_ISL_537470\|2020-04 | ···················· | ···················· | ···················· | ···················· | ···················· | ···················· |
| H | 2697 hCoV-19/USA/CA-UCLA-115/2020\|EPI_ISL_537471\|2020-0 | ···················· | ···················· | ···················· | ···················· | ···················· | ···················· |
| H | 2698 hCoV-19/USA/CA-UCLA-121/2020\|EPI_ISL_537475\|2020-0 | ···················· | ···················· | ···················· | ···················· | ···················· | ···················· |
| H | 2699 hCoV-19/USA/CA-UCLA-125/2020\|EPI_ISL_537476\|2020-0 | ···················· | ···················· | ···················· | ···················· | ···················· | ···················· |
| H | 2700 hCoV-19/USA/CA-UCLA-129/2020\|EPI_ISL_537477\|2020-0 | ···················· | ···················· | ···················· | ···················· | ···················· | ···················· |
| H | 2701 hCoV-19/USA/CA-UCLA-13/2020\|EPI_ISL_537478\|2020-04 | ···················· | ···················· | ···················· | ···················· | ···················· | ···················· |
| H | 2702 hCoV-19/USA/CA-UCLA-130/2020\|EPI_ISL_537479\|2020-0 | ···················· | ···················· | ···················· | ···················· | ···················· | ···················· |
| H | 2703 hCoV-19/USA/CA-UCLA-133/2020\|EPI_ISL_537480\|2020-0 | ···················· | ···················· | ···················· | ···················· | ···················· | ···················· |
| H | 2704 hCoV-19/USA/CA-UCLA-137/2020\|EPI_ISL_537481\|2020-0 | ···················· | ···················· | ···················· | ···················· | ···················· | ···················· |
| H | 2705 hCoV-19/USA/CA-UCLA-138/2020\|EPI_ISL_537482\|2020-0 | ···················· | ···················· | ···················· | ···················· | ···················· | ···················· |
| H | 2706 hCoV-19/USA/CA-UCLA-139/2020\|EPI_ISL_537483\|2020-0 | ···················· | ···················· | ···················· | ···················· | ···················· | ···················· |
| H | 2707 hCoV-19/USA/CA-UCLA-14/2020\|EPI_ISL_537484\|2020-04 | ···················· | ···················· | ···················· | ···················· | ···················· | ···················· |
| H | 2708 hCoV-19/USA/CA-UCLA-140/2020\|EPI_ISL_537485\|2020-0 | ···················· | ···················· | ···················· | ···················· | ···················· | ···················· |
| H | 2709 hCoV-19/USA/CA-UCLA-141/2020\|EPI_ISL_537486\|2020-0 | ···················· | ···················· | ···················· | ···················· | ···················· | ···················· |
| H | 2710 hCoV-19/USA/CA-UCLA-142/2020\|EPI_ISL_537487\|2020-0 | ···················· | ···················· | ···················· | ···················· | ···················· | ···················· |
| H | 2711 hCoV-19/USA/CA-UCLA-144/2020\|EPI_ISL_537488\|2020-0 | ···················· | ···················· | ···················· | ···················· | ···················· | ···················· |
| H | 2712 hCoV-19/USA/CA-UCLA-146/2020\|EPI_ISL_537489\|2020-0 | ···················· | ···················· | ···················· | ···················· | ···················· | ···················· |
| H | 2713 hCoV-19/USA/CA-UCLA-147/2020\|EPI_ISL_537490\|2020-0 | ···················· | ···················· | ···················· | ···················· | ···················· | ···················· |
| H | 2714 hCoV-19/USA/CA-UCLA-15/2020\|EPI_ISL_537491\|2020-04 | ···················· | ···················· | ···················· | ···················· | ···················· | ···················· |
| H | 2715 hCoV-19/USA/CA-UCLA-152/2020\|EPI_ISL_537492\|2020-0 | ···················· | ···················· | ···················· | ···················· | ···················· | ···················· |
| H | 2716 hCoV-19/USA/CA-UCLA-154/2020\|EPI_ISL_537493\|2020-0 | ···················· | ···················· | ···················· | ···················· | ···················· | ···················· |
| H | 2717 hCoV-19/USA/CA-UCLA-155/2020\|EPI_ISL_537494\|2020-0 | ···················· | ···················· | ···················· | ···················· | ···················· | ···················· |
| H | 2718 hCoV-19/USA/CA-UCLA-156/2020\|EPI_ISL_537495\|2020-0 | ···················· | ···················· | ···················· | ···················· | ···················· | ···················· |
| H | 2719 hCoV-19/USA/CA-UCLA-157/2020\|EPI_ISL_537496\|2020-0 | ···················· | ···················· | ···················· | ···················· | ···················· | ···················· |
| H | 2720 hCoV-19/USA/CA-UCLA-158/2020\|EPI_ISL_537497\|2020-0 | ···················· | ···················· | ···················· | ···················· | ···················· | ···················· |
| H | 2721 hCoV-19/USA/CA-UCLA-159/2020\|EPI_ISL_537498\|2020-0 | ···················· | ···················· | ···················· | ···················· | ···················· | ···················· |
| H | 2722 hCoV-19/USA/CA-UCLA-16/2020\|EPI_ISL_537499\|2020-04 | ···················· | ···················· | ···················· | ···················· | ···················· | ···················· |
| H | 2723 hCoV-19/England/LEED-2A9F19/2020\|EPI_ISL_538770\|20 | ···················· | ·A·················· | ···················· | ···················· | ···················· | ···················· |
| H | 2724 hCoV-19/England/LEED-2A976C/2020\|EPI_ISL_538772\|20 | ···················· | ···················· | ···················· | ···················· | ···················· | ···················· |
| H | 2725 hCoV-19/England/LEED-2A9DE2/2020\|EPI_ISL_538773\|20 | ···················· | ···················· | ···················· | ···················· | ···················· | ···················· |
| H | 2726 hCoV-19/England/LEED-2AA024/2020\|EPI_ISL_538774\|20 | ···················· | ···················· | ···················· | ···················· | ···················· | ···················· |
| H | 2727 hCoV-19/England/LEED-2A9702/2020\|EPI_ISL_538776\|20 | ···················· | ···················· | ···················· | ···················· | ···················· | ···················· |
| H | 2728 hCoV-19/England/LEED-2A9F0A/2020\|EPI_ISL_538777\|20 | ···················· | ·A·················· | ···················· | ···················· | ···················· | ···················· |
| H | 2729 hCoV-19/England/LEED-2A97E4/2020\|EPI_ISL_538778\|20 | ···················· | ···················· | ···················· | ···················· | ···················· | ···················· |
| H | 2730 hCoV-19/England/LEED-2A9EB2/2020\|EPI_ISL_538779\|20 | ···················· | ···················· | ···················· | ···················· | ···················· | ···················· |
| H | 2731 hCoV-19/England/LEED-2A9EFE/2020\|EPI_ISL_538780\|20 | ···················· | ···················· | ···················· | ···················· | ···················· | ···················· |

| | | | | | | | | | |
|---|---|---|---|---|---|---|---|---|---|
| H | 2732 | hCoV-19/England/LEED-2A9FDD/2020\|EPI_ISL_538781\|20 | .................... | .................... | .................... | .................... | .................... | .................... | |
| H | 2733 | hCoV-19/England/LEED-2A9FA0/2020\|EPI_ISL_538782\|20 | .................... | .................... | .................... | .................... | .................... | .................... | |
| H | 2734 | hCoV-19/England/LEED-2A9FBF/2020\|EPI_ISL_538783\|20 | .................... | .................... | .................... | .................... | .................... | .................... | |
| H | 2735 | hCoV-19/England/LEED-2A9F28/2020\|EPI_ISL_538784\|20 | .................... | .................... | .................... | .................... | .................... | .................... | |
| H | 2736 | hCoV-19/England/LEED-2AA19A/2020\|EPI_ISL_538786\|20 | .................... | .A.................. | .................... | .................... | .................... | .................... | |
| H | 2737 | hCoV-19/England/LEED-2AA22E/2020\|EPI_ISL_538787\|20 | .................... | .................... | .................... | .................... | .................... | .................... | |
| H | 2738 | hCoV-19/England/LEED-2A93FF/2020\|EPI_ISL_538788\|20 | .................... | .................... | .................... | .................... | .................... | .................... | |
| H | 2739 | hCoV-19/England/LEED-2AA419/2020\|EPI_ISL_538789\|20 | .................... | .................... | .................... | .................... | .................... | .................... | |
| H | 2740 | hCoV-19/England/LEED-2A9B8E/2020\|EPI_ISL_538790\|20 | .................... | .................... | .................... | .................... | .................... | .................... | |
| H | 2741 | hCoV-19/England/LEED-2AA491/2020\|EPI_ISL_538791\|20 | .................... | .................... | .................... | .................... | .................... | .................... | |
| H | 2742 | hCoV-19/England/LEED-2AA446/2020\|EPI_ISL_538792\|20 | .................... | .................... | .................... | .................... | .................... | .................... | |
| H | 2743 | hCoV-19/England/LEED-2A9580/2020\|EPI_ISL_538793\|20 | .................... | .A.................. | .................... | .................... | .................... | .................... | |
| H | 2744 | hCoV-19/England/LEED-2A940B/2020\|EPI_ISL_538794\|20 | .................... | .................... | .................... | .................... | .................... | .................... | |
| H | 2745 | hCoV-19/England/LEED-2AA4CE/2020\|EPI_ISL_538795\|20 | .................... | .................... | .................... | .................... | .................... | .................... | |
| H | 2746 | hCoV-19/England/LEED-2AA40A/2020\|EPI_ISL_538796\|20 | .................... | .................... | .........G.......... | .................... | .................... | .................... | |
| H | 2747 | hCoV-19/England/LEED-2AA525/2020\|EPI_ISL_538797\|20 | .................... | .................... | .................... | .................... | .................... | .................... | |
| H | 2748 | hCoV-19/England/LEED-2A9A36/2020\|EPI_ISL_538798\|20 | .................... | .................... | .................... | .................... | .................... | .................... | |
| H | 2749 | hCoV-19/England/LEED-2AA30D/2020\|EPI_ISL_538799\|20 | .................... | ...A................ | .................... | .................... | .................... | .................... | |
| H | 2750 | hCoV-19/England/QEUH-998426/2020\|EPI_ISL_539900\|20 | .................... | .................... | .................... | .................... | .................... | .................... | |
| H | 2751 | hCoV-19/England/QEUH-9983DE/2020\|EPI_ISL_539901\|20 | .................... | .................... | .................... | .................... | .................... | .................... | |
| H | 2752 | hCoV-19/England/QEUH-998471/2020\|EPI_ISL_539902\|20 | .................... | .................... | .................... | .................... | .................... | .................... | |
| H | 2753 | hCoV-19/England/QEUH-9987B4/2020\|EPI_ISL_539903\|20 | .................... | .................... | .................... | .................... | .................... | .................... | |
| H | 2754 | hCoV-19/England/QEUH-9985E7/2020\|EPI_ISL_539904\|20 | .................... | .................... | .................... | .................... | .................... | .................... | |
| H | 2755 | hCoV-19/Scotland/QEUH-9988DF/2020\|EPI_ISL_539905\|2 | .................... | .................... | .................... | .................... | .................... | .................... | |
| H | 2756 | hCoV-19/England/QEUH-99002A/2020\|EPI_ISL_539907\|20 | .................... | .................... | .................... | .................... | .................... | .................... | |
| H | 2757 | hCoV-19/England/QEUH-99882A/2020\|EPI_ISL_539908\|20 | .................... | .A.................. | .................... | .................... | .................... | .................... | |
| H | 2758 | hCoV-19/England/QEUH-9982E0/2020\|EPI_ISL_539909\|20 | .................... | .................... | .................... | .................... | .................... | .................... | |
| H | 2759 | hCoV-19/England/QEUH-99867B/2020\|EPI_ISL_539910\|20 | .................... | .................... | .................... | .................... | .................... | .................... | |
| H | 2760 | hCoV-19/England/QEUH-998CB5/2020\|EPI_ISL_539911\|20 | .................... | .................... | .................... | .................... | ...T................ | .................... | |
| H | 2761 | hCoV-19/England/QEUH-98F729/2020\|EPI_ISL_539912\|20 | .................... | .................... | .................... | .................... | .................... | .................... | |
| H | 2762 | hCoV-19/Scotland/QEUH-998532/2020\|EPI_ISL_539913\|2 | .................... | .................... | .................... | .................... | .................... | .................... | |
| H | 2763 | hCoV-19/England/QEUH-98FBD2/2020\|EPI_ISL_539914\|20 | .................... | .................... | .................... | .................... | .................... | .................... | |
| H | 2764 | hCoV-19/Scotland/QEUH-99857E/2020\|EPI_ISL_539915\|2 | .................... | .................... | .................... | .................... | .................... | .................... | |
| H | 2765 | hCoV-19/England/QEUH-9984DB/2020\|EPI_ISL_539916\|20 | .................... | .................... | .................... | .................... | .................... | .................... | |
| H | 2766 | hCoV-19/England/QEUH-98FCDF/2020\|EPI_ISL_539917\|20 | .................... | .................... | .................... | .................... | .................... | .................... | |
| H | 2767 | hCoV-19/England/QEUH-998B30/2020\|EPI_ISL_539918\|20 | .................... | .................... | .................... | .................... | .................... | .................... | |
| H | 2768 | hCoV-19/Scotland/QEUH-98F7A1/2020\|EPI_ISL_539919\|2 | .................... | .................... | .................... | .................... | .................... | .................... | |
| H | 2769 | hCoV-19/England/QEUH-99035E/2020\|EPI_ISL_539922\|20 | .................... | .................... | .................... | .................... | .................... | .................... | |
| H | 2770 | hCoV-19/England/QEUH-990163/2020\|EPI_ISL_539923\|20 | .................... | .................... | .................... | .................... | .................... | .................... | |
| H | 2771 | hCoV-19/England/QEUH-98FBC3/2020\|EPI_ISL_539924\|20 | .................... | .................... | .................... | .................... | .................... | .................... | |
| H | 2772 | hCoV-19/England/QEUH-990312/2020\|EPI_ISL_539925\|20 | .................... | .A.................. | .................... | .................... | .................... | ...A................ | |
| H | 2773 | hCoV-19/Wales/PHWC-16A513/2020\|EPI_ISL_540900\|2020 | .................... | .................... | .................... | .................... | .................... | .................... | |
| H | 2776 | hCoV-19/Wales/PHWC-16A540/2020\|EPI_ISL_540903\|2020 | .................... | .................... | .................... | .................... | .................... | .................... | |
| H | 2777 | hCoV-19/Wales/PHWC-16A55F/2020\|EPI_ISL_540904\|2020 | .................... | .................... | .................... | .................... | .................... | .................... | |
| H | 2778 | hCoV-19/Wales/PHWC-16A56E/2020\|EPI_ISL_540905\|2020 | .................... | .................... | .................... | .................... | .................... | .................... | |
| H | 2779 | hCoV-19/Wales/PHWC-16A57D/2020\|EPI_ISL_540906\|2020 | .................... | .................... | .................... | .................... | .................... | .................... | |
| H | 2780 | hCoV-19/Wales/PHWC-16A58C/2020\|EPI_ISL_540907\|2020 | .................... | .................... | .................... | .................... | .................... | .................... | |
| H | 2781 | hCoV-19/Wales/PHWC-16A59B/2020\|EPI_ISL_540908\|2020 | .................... | .................... | .................... | .................... | .................... | .................... | |
| H | 2782 | hCoV-19/Wales/PHWC-16A5AA/2020\|EPI_ISL_540909\|2020 | .................... | .................... | .................... | .................... | .................... | .................... | |
| H | 2783 | hCoV-19/USA/TX-HMH-3748/2020\|EPI_ISL_543160\|2020-0 | .................... | .................... | .................... | .................... | .................... | N.....NN............ | 1 |
| H | 2784 | hCoV-19/USA/TX-HMH-3750/2020\|EPI_ISL_543161\|2020-0 | .................... | .................... | .................... | .................... | .............N...... | ......NN............ | |
| H | 2785 | hCoV-19/USA/TX-HMH-3753/2020\|EPI_ISL_543164\|2020-0 | .................... | .................... | .................... | .................... | .................... | .................... | |
| H | 2786 | hCoV-19/USA/TX-HMH-3754/2020\|EPI_ISL_543165\|2020-0 | .................... | .................... | .................... | .................... | .................... | .................... | |
| H | 2787 | hCoV-19/USA/TX-HMH-3755/2020\|EPI_ISL_543166\|2020-0 | .................... | .................... | .................... | .................... | .................... | .................... | |
| H | 2788 | hCoV-19/USA/TX-HMH-3758/2020\|EPI_ISL_543169\|2020-0 | .................... | .................... | .................... | .................... | .................... | .................... | |
| H | 2804 | hCoV-19/USA/TX-HMH-3784/2020\|EPI_ISL_543189\|2020-0 | .................... | .................... | .................... | .................... | .................... | .................... | |
| H | 2806 | hCoV-19/USA/TX-HMH-4019/2020\|EPI_ISL_543301\|2020-0 | .................... | .................... | .................... | .................... | .................... | .................... | |
| H | 2807 | hCoV-19/USA/TX-HMH-4021/2020\|EPI_ISL_543303\|2020-0 | .................... | .................... | .................... | .................... | .................... | .................... | |
| H | 2808 | hCoV-19/USA/TX-HMH-4022/2020\|EPI_ISL_543304\|2020-0 | .................... | .................... | .................... | .................... | .................... | .................... | |
| H | 2809 | hCoV-19/USA/TX-HMH-4023/2020\|EPI_ISL_543305\|2020-0 | .................... | .................... | .................... | .................... | .................... | .................... | |
| H | 2810 | hCoV-19/USA/TX-HMH-4024/2020\|EPI_ISL_543306\|2020-0 | .................... | .................... | .................... | .................... | .................... | .................... | |
| H | 2811 | hCoV-19/USA/TX-HMH-4028/2020\|EPI_ISL_543308\|2020-0 | .................... | .................... | .................... | .................... | .................... | .................... | |
| H | 2813 | hCoV-19/USA/TX-HMH-4031/2020\|EPI_ISL_543311\|2020-0 | .................... | .................... | .................... | .................... | .................... | .................... | |
| H | 2814 | hCoV-19/USA/TX-HMH-4032/2020\|EPI_ISL_543312\|2020-0 | .................... | ............A....... | .................... | .................... | .................... | .................... | |
| H | 2816 | hCoV-19/Peru/LIM-INS-001/2020\|EPI_ISL_545955\|2020- | .................... | .................... | .................... | .................... | .................... | .................... | |
| H | 2817 | hCoV-19/Scotland/QEUH-9698C8/2020\|EPI_ISL_531100\|2 | .................... | .................... | .................... | .................... | .................... | .................... | |
| H | 2818 | hCoV-19/England/QEUH-9651D0/2020\|EPI_ISL_531101\|20 | .................... | .................... | .................... | .................... | .................... | .................... | |
| H | 2819 | hCoV-19/Wales/QEUH-969AC2/2020\|EPI_ISL_531102\|2020 | .................... | .................... | .................... | .................... | .................... | .................... | |
| H | 2820 | hCoV-19/Scotland/QEUH-964F22/2020\|EPI_ISL_531103\|2 | .................... | .................... | .................... | .................... | .................... | .................... | |
| H | 2821 | hCoV-19/Scotland/QEUH-965589/2020\|EPI_ISL_531104\|2 | .................... | .................... | .................... | .................... | .................... | .................... | |
| H | 2822 | hCoV-19/Northern Ireland/QEUH-969A59/2020\|EPI_ISL_ | .................... | .................... | .................... | .................... | .................... | .................... | |
| H | 2823 | hCoV-19/England/QEUH-96598D/2020\|EPI_ISL_531106\|20 | .................... | .................... | .................... | .................... | .................... | .................... | |
| H | 2824 | hCoV-19/Northern Ireland/QEUH-969813/2020\|EPI_ISL_ | .................... | .................... | .................... | .................... | .................... | .................... | |
| H | 2825 | hCoV-19/England/QEUH-969567/2020\|EPI_ISL_531108\|20 | .................... | .A.................. | .................... | .................... | .................... | ...A................ | |
| H | 2826 | hCoV-19/England/QEUH-9652EC/2020\|EPI_ISL_531109\|20 | .................... | .................... | .................... | .................... | .................... | .................... | |
| H | 2827 | hCoV-19/Northern Ireland/QEUH-965431/2020\|EPI_ISL_ | .................... | .................... | .................... | .................... | .................... | .................... | |
| H | 2828 | hCoV-19/England/QEUH-964C49/2020\|EPI_ISL_531112\|20 | .................... | .................... | .................... | .................... | .................... | .................... | |
| H | 2829 | hCoV-19/England/QEUH-9695FE/2020\|EPI_ISL_531113\|20 | .................... | .................... | .................... | .................... | .................... | .................... | |
| H | 2830 | hCoV-19/Scotland/QEUH-9655E3/2020\|EPI_ISL_531116\|2 | .................... | .................... | .................... | .................... | .................... | .................... | |
| H | 2831 | hCoV-19/England/QEUH-965880/2020\|EPI_ISL_531117\|20 | .................... | .................... | .................... | .................... | .................... | .................... | |

| | | | | | | | | |
|---|---|---|---|---|---|---|---|---|
| H | 2832 | hCoV-19/Scotland/QEUH-969822/2020\|EPI_ISL_531118\|2 | ···················· | ···················· | ····················· | ···················· | ···················· | ···················· |
| H | 2833 | hCoV-19/England/QEUH-965695/2020\|EPI_ISL_531119\|20 | ···················· | ···················· | ····················· | ···················· | ···················· | ···················· |
| H | 2834 | hCoV-19/Northern Ireland/QEUH-96995C/2020\|EPI_ISL_ | ···················· | ···················· | ····················· | ···················· | ···················· | ···················· |
| H | 2835 | hCoV-19/Northern Ireland/QEUH-964CEF/2020\|EPI_ISL_ | ···················· | ···················· | ····················· | ···················· | ···················· | ···················· |
| H | 2836 | hCoV-19/Scotland/QEUH-964960/2020\|EPI_ISL_531123\|2 | ···················· | ···················· | ····················· | ···················· | ···················· | ···················· |
| H | 2837 | hCoV-19/England/QEUH-96499D/2020\|EPI_ISL_531124\|20 | ···················· | ·A·················· | ····················· | ···················· | ···················· | ··A················· |
| H | 2838 | hCoV-19/England/QEUH-964D73/2020\|EPI_ISL_531125\|20 | ···················· | ···················· | ····················· | ···················· | ···················· | ···················· |
| H | 2839 | hCoV-19/Scotland/QEUH-9654C8/2020\|EPI_ISL_531126\|2 | ···················· | ···················· | ····················· | ···················· | ···················· | ···················· |
| H | 2840 | hCoV-19/Wales/QEUH-9648DC/2020\|EPI_ISL_531127\|2020 | ···················· | ···················· | ····················· | ···················· | ···················· | ···················· |
| H | 2841 | hCoV-19/Northern Ireland/QEUH-964E70/2020\|EPI_ISL_ | ···················· | ···················· | ····················· | ···················· | ···················· | ···················· |
| H | 2842 | hCoV-19/England/QEUH-969691/2020\|EPI_ISL_531129\|20 | ···················· | ···················· | ····················· | ···················· | ···················· | ···················· |
| H | 2843 | hCoV-19/Scotland/QEUH-964E61/2020\|EPI_ISL_531130\|2 | ···················· | ···················· | ····················· | ···················· | ···················· | ···················· |
| H | 2844 | hCoV-19/England/QEUH-9651FE/2020\|EPI_ISL_531133\|20 | ···················· | ·A·················· | ····················· | ···················· | ···················· | ··A················· |
| H | 2845 | hCoV-19/Northern Ireland/QEUH-965194/2020\|EPI_ISL_ | ···················· | ···················· | ····················· | ···················· | ···················· | ···················· |
| H | 2846 | hCoV-19/England/QEUH-9699B6/2020\|EPI_ISL_531135\|20 | ···················· | ···················· | ····················· | ···················· | ···················· | ···················· |
| H | 2847 | hCoV-19/England/QEUH-9652CE/2020\|EPI_ISL_531136\|20 | ···················· | ·A·················· | ····················· | ···················· | ···················· | ··A················· |
| H | 2848 | hCoV-19/England/QEUH-9694D3/2020\|EPI_ISL_531137\|20 | ···················· | ·A·················· | ····················· | ···················· | ···················· | ··A················· |
| H | 2849 | hCoV-19/Wales/QEUH-969B29/2020\|EPI_ISL_531138\|2020 | ···················· | ···················· | ····················· | ···················· | ···················· | ···················· |
| H | 2850 | hCoV-19/England/QEUH-969628/2020\|EPI_ISL_531139\|20 | ···················· | ···················· | ····················· | ···················· | ···················· | ···················· |
| H | 2851 | hCoV-19/England/QEUH-965273/2020\|EPI_ISL_531140\|20 | ···················· | ···················· | ····················· | ···················· | ···················· | ···················· |
| H | 2852 | hCoV-19/England/QEUH-9648FA/2020\|EPI_ISL_531141\|20 | ···················· | ···················· | ····················· | ···················· | ···················· | ···················· |
| H | 2853 | hCoV-19/Scotland/QEUH-964951/2020\|EPI_ISL_531142\|2 | ···················· | ···················· | ····················· | ···················· | ···················· | ···················· |
| H | 2854 | hCoV-19/England/QEUH-9694C4/2020\|EPI_ISL_531145\|20 | ···················· | ·A·················· | ····················· | ···················· | ···················· | ··A················· |
| H | 2855 | hCoV-19/England/QEUH-964DA0/2020\|EPI_ISL_531146\|20 | ···················· | ···················· | ····················· | ···················· | ···················· | ···················· |
| H | 2856 | hCoV-19/Scotland/QEUH-9654E6/2020\|EPI_ISL_531147\|2 | ···················· | ···················· | ····················· | ···················· | ···················· | ···················· |
| H | 2857 | hCoV-19/England/QEUH-96571A/2020\|EPI_ISL_531150\|20 | ···················· | ···················· | ····················· | ···················· | ···················· | ···················· |
| H | 2858 | hCoV-19/England/QEUH-964906/2020\|EPI_ISL_531151\|20 | ···················· | ···················· | ····················· | ···················· | ···················· | ···················· |
| H | 2859 | hCoV-19/England/QEUH-964AE5/2020\|EPI_ISL_531152\|20 | ···················· | ···················· | ····················· | ···················· | ···················· | ···················· |
| H | 2860 | hCoV-19/Scotland/QEUH-96506A/2020\|EPI_ISL_531154\|2 | ···················· | ···················· | ····················· | ···················· | ···················· | ···················· |
| H | 2861 | hCoV-19/Scotland/QEUH-9657A1/2020\|EPI_ISL_531155\|2 | ···················· | ···················· | ····················· | ···················· | ···················· | ···················· |
| H | 2862 | hCoV-19/Scotland/QEUH-964F8C/2020\|EPI_ISL_531156\|2 | ···················· | ···················· | ····················· | ···················· | ···················· | ···················· |
| H | 2863 | hCoV-19/Scotland/QEUH-964AF4/2020\|EPI_ISL_531157\|2 | ···················· | ···················· | ····················· | ···················· | ···················· | ···················· |
| H | 2864 | hCoV-19/England/QEUH-969BA1/2020\|EPI_ISL_531158\|20 | ···················· | ···················· | ····················· | ···················· | ···················· | ···················· |
| H | 2865 | hCoV-19/England/QEUH-95F50A/2020\|EPI_ISL_532424\|20 | ···················· | ···················· | ····················· | ···················· | ···················· | ···················· |
| H | 2866 | hCoV-19/England/QEUH-95F564/2020\|EPI_ISL_532425\|20 | ···················· | ···················· | ····················· | ···················· | ···················· | ···················· |
| H | 2867 | hCoV-19/England/QEUH-95F8E3/2020\|EPI_ISL_532426\|20 | ···················· | ···················· | ····················· | ···················· | ···················· | ···················· |
| H | 2868 | hCoV-19/England/QEUH-95F704/2020\|EPI_ISL_532428\|20 | ···················· | ···················· | ····················· | ···················· | ···················· | ···················· |
| H | 2869 | hCoV-19/England/QEUH-95F212/2020\|EPI_ISL_532429\|20 | ···················· | ···················· | ····················· | ···················· | ···················· | ···················· |
| H | 2870 | hCoV-19/Scotland/QEUH-95EDA9/2020\|EPI_ISL_532430\|2 | ···················· | ···················· | ····················· | ···················· | ···················· | ···················· |
| H | 2871 | hCoV-19/Scotland/QEUH-95F397/2020\|EPI_ISL_532432\|2 | ···················· | ···················· | ····················· | ···················· | ···················· | ···················· |
| H | 2872 | hCoV-19/England/QEUH-95FA74/2020\|EPI_ISL_532433\|20 | ···················· | ···················· | ····················· | ···················· | ···················· | ···················· |
| H | 2873 | hCoV-19/England/QEUH-95F230/2020\|EPI_ISL_532436\|20 | ···················· | ···················· | ····················· | ···················· | ···················· | ···················· |
| H | 2874 | hCoV-19/England/QEUH-95F582/2020\|EPI_ISL_532437\|20 | ···················· | ···················· | ····················· | ···················· | ···················· | ···················· |
| H | 2875 | hCoV-19/England/QEUH-95F898/2020\|EPI_ISL_532439\|20 | ···················· | ···················· | ····················· | ···················· | ···················· | ···················· |
| H | 2876 | hCoV-19/Scotland/QEUH-95E8A8/2020\|EPI_ISL_532440\|2 | ···················· | ···················· | ····················· | ···················· | ···················· | ···················· |
| H | 2877 | hCoV-19/England/QEUH-95F8D4/2020\|EPI_ISL_532442\|20 | ···················· | ···················· | ····················· | ···················· | ···················· | ···················· |
| H | 2878 | hCoV-19/Scotland/QEUH-95F41C/2020\|EPI_ISL_532444\|2 | ···················· | ···················· | ····················· | ···················· | ···················· | ···················· |
| H | 2879 | hCoV-19/Scotland/QEUH-95F731/2020\|EPI_ISL_532446\|2 | ···················· | ···················· | ····················· | ···················· | ···················· | ···················· |
| H | 2880 | hCoV-19/Northern Ireland/QEUH-95ED5E/2020\|EPI_ISL_ | ···················· | ···················· | ····················· | ···················· | ···················· | ···················· |
| H | 2881 | hCoV-19/England/QEUH-95FC41/2020\|EPI_ISL_532449\|20 | ···················· | ···················· | ····················· | ···················· | ···················· | ···················· |
| H | 2882 | hCoV-19/Northern Ireland/QEUH-95EDE5/2020\|EPI_ISL_ | ···················· | ···················· | ····················· | ···················· | ···················· | ···················· |
| H | 2883 | hCoV-19/Northern Ireland/QEUH-95FADE/2020\|EPI_ISL_ | ···················· | ···················· | ····················· | ···················· | ···················· | ···················· |
| H | 2884 | hCoV-19/Scotland/QEUH-95ED03/2020\|EPI_ISL_532452\|2 | ···················· | ···················· | ····················· | ···················· | ···················· | ···················· |
| H | 2885 | hCoV-19/England/QEUH-95E93C/2020\|EPI_ISL_532453\|20 | ···················· | ·A·················· | ····················· | ···················· | ···················· | ··A················· |
| H | 2886 | hCoV-19/Scotland/QEUH-95E653/2020\|EPI_ISL_532454\|2 | ···················· | ···················· | ····················· | ···················· | ···················· | ···················· |
| H | 2887 | hCoV-19/England/QEUH-95FB35/2020\|EPI_ISL_532455\|20 | ···················· | ···················· | ····················· | ···················· | ···················· | ···················· |
| H | 2888 | hCoV-19/Scotland/QEUH-95F713/2020\|EPI_ISL_532456\|2 | ···················· | ···················· | ····················· | ···················· | ···················· | ···················· |
| H | 2889 | hCoV-19/England/QEUH-95ECD9/2020\|EPI_ISL_532457\|20 | ···················· | ···················· | ····················· | ···················· | ···················· | ···················· |
| H | 2890 | hCoV-19/England/QEUH-95F26D/2020\|EPI_ISL_532458\|20 | ···················· | ···················· | ····················· | ···················· | ···················· | ···················· |
| H | 2891 | hCoV-19/England/QEUH-95E608/2020\|EPI_ISL_532459\|20 | ···················· | ···················· | ····················· | ···················· | ···················· | ···················· |
| H | 2892 | hCoV-19/England/QEUH-95F3F1/2020\|EPI_ISL_532461\|20 | ···················· | ···················· | ····················· | ···················· | ···················· | ···················· |
| H | 2893 | hCoV-19/England/QEUH-95E8E4/2020\|EPI_ISL_532462\|20 | ···················· | ···················· | ····················· | ···················· | ···················· | ···················· |
| H | 2894 | hCoV-19/Scotland/QEUH-95F616/2020\|EPI_ISL_532463\|2 | ···················· | ···················· | ····················· | ···················· | ···················· | ···················· |
| H | 2895 | hCoV-19/Scotland/QEUH-95F33D/2020\|EPI_ISL_532464\|2 | ···················· | ···················· | ····················· | ···················· | ···················· | ···················· |
| H | 2896 | hCoV-19/England/QEUH-95E5B0/2020\|EPI_ISL_532465\|20 | ···················· | ···················· | ····················· | ···················· | ···················· | ···················· |
| H | 2897 | hCoV-19/England/QEUH-95F24F/2020\|EPI_ISL_532466\|20 | ···················· | ···················· | ····················· | ···················· | ···················· | ···················· |
| H | 2898 | hCoV-19/England/QEUH-95F5CE/2020\|EPI_ISL_532467\|20 | ···················· | ···················· | ····················· | ···················· | ···················· | ···················· |
| H | 2899 | hCoV-19/Northern Ireland/QEUH-95EBBE/2020\|EPI_ISL_ | ···················· | ···················· | ····················· | ···················· | ···················· | ···················· |
| H | 2900 | hCoV-19/England/QEUH-95EF1C/2020\|EPI_ISL_532470\|20 | ···················· | ···················· | ····················· | ···················· | ···················· | ···················· |
| H | 2901 | hCoV-19/England/QEUH-95E83F/2020\|EPI_ISL_532471\|20 | ···················· | ···················· | ····················· | ···················· | ···················· | ···················· |
| H | 2902 | hCoV-19/Scotland/QEUH-95F6AD/2020\|EPI_ISL_532472\|2 | ···················· | ···················· | ····················· | ···················· | ···················· | ···················· |
| H | 2903 | hCoV-19/Scotland/QEUH-961EDD/2020\|EPI_ISL_532473\|2 | ···················· | ···················· | ····················· | ···················· | ···················· | ···················· |
| H | 2904 | hCoV-19/Northern Ireland/QEUH-95EC51/2020\|EPI_ISL_ | ···················· | ···················· | ····················· | ···················· | ···················· | ···················· |
| H | 2905 | hCoV-19/Northern Ireland/QEUH-95EED3/2020\|EPI_ISL_ | ···················· | ···················· | ····················· | ···················· | ···················· | ···················· |
| H | 2906 | hCoV-19/England/QEUH-95FB53/2020\|EPI_ISL_532477\|20 | ···················· | ···················· | ····················· | ···················· | ···················· | ···················· |
| H | 2907 | hCoV-19/England/QEUH-95ED7C/2020\|EPI_ISL_532478\|20 | ···················· | ···················· | ····················· | ···················· | ···················· | ···················· |
| H | 2908 | hCoV-19/Scotland/QEUH-95ED6D/2020\|EPI_ISL_532480\|2 | ···················· | ···················· | ····················· | ···················· | ···················· | ···················· |
| H | 2909 | hCoV-19/Northern Ireland/QEUH-95E84E/2020\|EPI_ISL_ | ···················· | ···················· | ····················· | ···················· | ···················· | ···················· |
| H | 2910 | hCoV-19/Northern Ireland/QEUH-95E9F0/2020\|EPI_ISL_ | ···················· | ·A·················· | ····················· | ···················· | ···················· | ···················· |
| H | 2911 | hCoV-19/England/QEUH-95FACF/2020\|EPI_ISL_532483\|20 | ···················· | ···················· | ····················· | ···················· | ···················· | ···················· |

| | | | | | | | | |
|---|---|---|---|---|---|---|---|---|
| H | 2912 | hCoV-19/Scotland/QEUH-95E8B7/2020\|EPI_ISL_532484\|2 | ···················· | ···················· | ····················· | ···················· | ···················· | ···················· |
| H | 2913 | hCoV-19/England/QEUH-95FB44/2020\|EPI_ISL_532485\|20 | ···················· | ···················· | ····················· | ···················· | ···················· | ···················· |
| H | 2914 | hCoV-19/England/QEUH-95FAB0/2020\|EPI_ISL_532487\|20 | ···················· | ···················· | ····················· | ···················· | ···················· | ···················· |
| H | 2915 | hCoV-19/England/QEUH-95E91E/2020\|EPI_ISL_532488\|20 | ···················· | ···················· | ····················· | ···················· | ···················· | ···················· |
| H | 2916 | hCoV-19/England/QEUH-95F142/2020\|EPI_ISL_532489\|20 | ···················· | ·A·······A·········· | ····················· | ···················· | ···················· | ···················· |
| H | 2930 | hCoV-19/Spain/CL-IBV-97006166/2020\|EPI_ISL_537313\| | ···················· | ···················· | ····················· | ···················· | ···················· | ···················· |
| H | 2935 | hCoV-19/England/QEUH-98FFE5/2020\|EPI_ISL_539921\|20 | ···················· | ···················· | ····················· | ···················· | ···················· | ···················· |
| H | 2936 | hCoV-19/England/QEUH-998E64/2020\|EPI_ISL_539926\|20 | ···················· | ···················· | ····················· | ···················· | ···················· | ···················· |
| H | 2937 | hCoV-19/England/QEUH-998BD6/2020\|EPI_ISL_539927\|20 | ···················· | ···················· | ····················· | ···················· | ···················· | ···················· |
| H | 2938 | hCoV-19/Scotland/QEUH-98FB4B/2020\|EPI_ISL_539928\|2 | ···················· | ···················· | ····················· | ···················· | ···················· | ···················· |
| H | 2939 | hCoV-19/England/QEUH-98F62C/2020\|EPI_ISL_539929\|20 | ···················· | ···················· | ····················· | ···················· | ···················· | ···················· |
| H | 2940 | hCoV-19/England/QEUH-98FCEE/2020\|EPI_ISL_539930\|20 | ···················· | ···················· | ····················· | ···················· | ···················· | ···················· |
| H | 2941 | hCoV-19/England/QEUH-9982C2/2020\|EPI_ISL_539931\|20 | ···················· | ···················· | ····················· | ···················· | ···················· | ···················· |
| H | 2942 | hCoV-19/England/QEUH-9985D8/2020\|EPI_ISL_539932\|20 | ···················· | ·A·················· | ····················· | ···················· | ···················· | ···················· |
| H | 2943 | hCoV-19/England/QEUH-9983FC/2020\|EPI_ISL_539933\|20 | ···················· | ·A·················· | ····················· | ···················· | ···················· | ···················· |
| H | 2944 | hCoV-19/England/QEUH-98FB3C/2020\|EPI_ISL_539934\|20 | ···················· | ···················· | ····················· | ···················· | ···················· | ···················· |
| H | 2945 | hCoV-19/Scotland/QEUH-98F9E7/2020\|EPI_ISL_539935\|2 | ···················· | ···A················ | ····················· | ···················· | ···················· | ···················· |
| H | 2946 | hCoV-19/England/QEUH-98FF21/2020\|EPI_ISL_539936\|20 | ···················· | ···················· | ···········T········· | ···················· | ···················· | ···················· |
| H | 2947 | hCoV-19/England/QEUH-98FA02/2020\|EPI_ISL_539937\|20 | ···················· | ···················· | ····················· | ···················· | ······T············· | ···················· |
| H | 2948 | hCoV-19/England/QEUH-9901EB/2020\|EPI_ISL_539938\|20 | ···················· | ·A·················· | ····················· | ···················· | ···················· | ···A················ |
| H | 2949 | hCoV-19/Scotland/QEUH-9900B1/2020\|EPI_ISL_539939\|2 | ···················· | ···················· | ····················· | ···················· | ···················· | ···················· |
| H | 2950 | hCoV-19/England/QEUH-98FD90/2020\|EPI_ISL_539941\|20 | ···················· | ·A·················· | ····················· | ···················· | ···················· | ···················· |
| H | 2951 | hCoV-19/England/QEUH-9988C0/2020\|EPI_ISL_539942\|20 | ···················· | ···················· | ····················· | ···················· | ···················· | ···················· |
| H | 2952 | hCoV-19/England/QEUH-998AF7/2020\|EPI_ISL_539943\|20 | ···················· | ···················· | ····················· | ···················· | ···················· | ···················· |
| H | 2953 | hCoV-19/Scotland/QEUH-998936/2020\|EPI_ISL_539944\|2 | ···················· | ···················· | ····················· | ···················· | ···················· | ···················· |
| H | 2954 | hCoV-19/England/QEUH-99880C/2020\|EPI_ISL_539945\|20 | ···················· | ···················· | ····················· | ···················· | ···················· | ···················· |
| H | 2955 | hCoV-19/England/QEUH-98FF30/2020\|EPI_ISL_539946\|20 | ···················· | ···················· | ····················· | ···················· | ···················· | ···················· |
| H | 2956 | hCoV-19/England/QEUH-9984AE/2020\|EPI_ISL_539947\|20 | ···················· | ···················· | ····················· | ···················· | ···················· | ···················· |
| H | 2957 | hCoV-19/England/QEUH-98FE06/2020\|EPI_ISL_539948\|20 | ···················· | ···················· | ····················· | ···················· | ···················· | ···················· |
| H | 2958 | hCoV-19/England/QEUH-998B4F/2020\|EPI_ISL_539949\|20 | ···················· | ···················· | ····················· | ···················· | ···················· | ···················· |
| H | 2959 | hCoV-19/England/QEUH-98FE60/2020\|EPI_ISL_539950\|20 | ···················· | ···················· | ····················· | ···················· | ···················· | ···················· |
| H | 2960 | hCoV-19/England/QEUH-998848/2020\|EPI_ISL_539951\|20 | ···················· | ···················· | ····················· | ···················· | ···················· | ···················· |
| H | 2961 | hCoV-19/England/QEUH-998C79/2020\|EPI_ISL_539952\|20 | ···················· | ·A·················· | ····················· | ···················· | ···················· | ···················· |
| H | 2962 | hCoV-19/England/QEUH-98F47D/2020\|EPI_ISL_539953\|20 | ···················· | ·A·················· | ····················· | ···················· | ···················· | ···················· |
| H | 2963 | hCoV-19/England/QEUH-9983ED/2020\|EPI_ISL_539954\|20 | ···················· | ···················· | ····················· | ···················· | ···················· | ···················· |
| H | 2964 | hCoV-19/Scotland/QEUH-998A06/2020\|EPI_ISL_539955\|2 | ···················· | ···················· | ····················· | ···················· | ···················· | ···················· |
| H | 2965 | hCoV-19/England/QEUH-98F826/2020\|EPI_ISL_539956\|20 | ···················· | ···················· | ····················· | ···················· | ···················· | ···················· |
| H | 2966 | hCoV-19/Scotland/QEUH-990172/2020\|EPI_ISL_539957\|2 | ···················· | ···················· | ····················· | ···················· | ···················· | ···················· |
| H | 2967 | hCoV-19/England/QEUH-9982FF/2020\|EPI_ISL_539958\|20 | ···················· | ···················· | ····················· | ···················· | ···················· | ···················· |
| H | 2968 | hCoV-19/Scotland/QEUH-98F61D/2020\|EPI_ISL_539959\|2 | ···················· | ···················· | ····················· | ···················· | ···················· | ···················· |
| H | 2969 | hCoV-19/Scotland/QEUH-9982D1/2020\|EPI_ISL_539960\|2 | ···················· | ···················· | ····················· | ···················· | ···················· | ···················· |
| H | 2970 | hCoV-19/England/QEUH-9901DC/2020\|EPI_ISL_539964\|20 | ···················· | ···················· | ····················· | ···················· | ···················· | ···················· |
| H | 2971 | hCoV-19/England/QEUH-99868A/2020\|EPI_ISL_539965\|20 | ···················· | ·A·················· | ····················· | ···················· | ···················· | ···················· |
| H | 2972 | hCoV-19/England/QEUH-9902F7/2020\|EPI_ISL_539966\|20 | ···················· | ···················· | ····················· | ···················· | ···················· | ···················· |
| H | 2973 | hCoV-19/England/QEUH-99830B/2020\|EPI_ISL_539967\|20 | ···················· | ···················· | ····················· | ···················· | ···················· | ···················· |
| H | 2974 | hCoV-19/England/QEUH-9989BE/2020\|EPI_ISL_539969\|20 | ···················· | ···················· | ····················· | ···················· | ···················· | ···················· |
| H | 2975 | hCoV-19/England/QEUH-998444/2020\|EPI_ISL_539970\|20 | ···················· | ···················· | ····················· | ···················· | ···················· | ···················· |
| H | 2976 | hCoV-19/England/QEUH-98FAE4/2020\|EPI_ISL_539971\|20 | ···················· | ···················· | ····················· | ···················· | ···················· | ···················· |
| H | 2977 | hCoV-19/England/QEUH-998B5E/2020\|EPI_ISL_539972\|20 | ···················· | ···················· | ····················· | ···················· | ···················· | ···················· |
| H | 2978 | hCoV-19/England/QEUH-98FDEB/2020\|EPI_ISL_539973\|20 | ···················· | ···················· | ····················· | ···················· | ···················· | ···················· |
| H | 2979 | hCoV-19/England/QEUH-98FB1E/2020\|EPI_ISL_539974\|20 | ···················· | ···················· | ····················· | ···················· | ···················· | ···················· |
| H | 2980 | hCoV-19/England/QEUH-98F49B/2020\|EPI_ISL_539975\|20 | ···················· | ···················· | ····················· | ···················· | ···················· | ···················· |
| H | 2981 | hCoV-19/England/QEUH-998C4C/2020\|EPI_ISL_539976\|20 | ···················· | ···················· | ····················· | ···················· | ···················· | ···················· |
| H | 2982 | hCoV-19/England/QEUH-990066/2020\|EPI_ISL_539977\|20 | ···················· | ···················· | ····················· | ···················· | ···················· | ···················· |
| H | 2983 | hCoV-19/England/QEUH-998E28/2020\|EPI_ISL_539978\|20 | ···················· | ·A·················· | ····················· | ···················· | ···················· | ···················· |
| H | 2984 | hCoV-19/England/QEUH-990303/2020\|EPI_ISL_539979\|20 | ···················· | ···················· | ····················· | ···················· | ···················· | ···················· |
| H | 2985 | hCoV-19/Wales/PHWC-16A5B9/2020\|EPI_ISL_540910\|2020 | ···················· | ···················· | ····················· | ···················· | ···················· | ···················· |
| H | 2986 | hCoV-19/Wales/PHWC-16A5C8/2020\|EPI_ISL_540911\|2020 | ···················· | ···················· | ····················· | ···················· | ···················· | ···················· |
| H | 2987 | hCoV-19/Wales/PHWC-16A5D7/2020\|EPI_ISL_540912\|2020 | ···················· | ···················· | ····················· | ···················· | ···················· | ···················· |
| H | 2988 | hCoV-19/Wales/PHWC-16A5E6/2020\|EPI_ISL_540913\|2020 | ···················· | ···················· | ····················· | ···················· | ···················· | ···················· |
| H | 2989 | hCoV-19/Wales/PHWC-16A5F5/2020\|EPI_ISL_540914\|2020 | ···················· | ···················· | ····················· | ···················· | ···················· | ···················· |
| H | 2990 | hCoV-19/Wales/PHWC-16A601/2020\|EPI_ISL_540915\|2020 | ···················· | ···················· | ····················· | ···················· | ···················· | ···················· |
| H | 2991 | hCoV-19/Wales/PHWC-16A610/2020\|EPI_ISL_540916\|2020 | ···················· | ···················· | ····················· | ···················· | ···················· | ···················· |
| H | 2992 | hCoV-19/Wales/PHWC-16A62F/2020\|EPI_ISL_540917\|2020 | ···················· | ···················· | ····················· | ···················· | ···················· | ···················· |
| H | 2993 | hCoV-19/Wales/PHWC-16A63E/2020\|EPI_ISL_540918\|2020 | ···················· | ···················· | ····················· | ···················· | ···················· | ···················· |
| H | 2994 | hCoV-19/Wales/PHWC-16A64D/2020\|EPI_ISL_540919\|2020 | ···················· | ···················· | ····················· | ···················· | ···················· | ···················· |
| H | 2995 | hCoV-19/Wales/PHWC-16A65C/2020\|EPI_ISL_540920\|2020 | ···················· | ···················· | ····················· | ···················· | ···················· | ···················· |
| H | 2996 | hCoV-19/USA/WY-WYPHL-20015627/2020\|EPI_ISL_540921\| | ···················· | ···················· | ····················· | ···················· | ···················· | ···················· |
| H | 2997 | hCoV-19/Peru/ANC-UPCH-0010/2020\|EPI_ISL_540923\|202 | ···················· | ···················· | ····················· | ···················· | ···················· | ···················· |
| H | 2998 | hCoV-19/Peru/ARE-UPCH-0016/2020\|EPI_ISL_540924\|202 | ···················· | ···················· | ····················· | ···················· | ···················· | ···················· |
| H | 2999 | hCoV-19/Peru/LIM-UPCH-0024/2020\|EPI_ISL_540925\|202 | ···················· | ···················· | ····················· | ···················· | ···················· | ···················· |
| H | 3000 | hCoV-19/Peru/LIM-UPCH-0025/2020\|EPI_ISL_540926\|202 | ···················· | ···················· | ····················· | ···················· | ···················· | ···················· |
| H | 3001 | hCoV-19/Peru/CAL-UPCH-0026/2020\|EPI_ISL_540927\|202 | ···················· | ···················· | ····················· | ···················· | ···················· | ···················· |
| H | 3002 | hCoV-19/Peru/CAL-UPCH-0027/2020\|EPI_ISL_540928\|202 | ···················· | ···················· | ····················· | ···················· | ···················· | ···················· |
| H | 3003 | hCoV-19/Peru/LIM-UPCH-0029/2020\|EPI_ISL_540929\|202 | ···················· | ···················· | ····················· | ···················· | ···················· | ···················· |
| H | 3004 | hCoV-19/Peru/LIM-UPCH-0030/2020\|EPI_ISL_540930\|202 | ···················· | ···················· | ····················· | ···················· | ···················· | ···················· |
| H | 3005 | hCoV-19/Peru/LIM-UPCH-0031/2020\|EPI_ISL_540931\|202 | ···················· | ···················· | ····················· | ···················· | ···················· | ···················· |
| H | 3006 | hCoV-19/Peru/LIM-UPCH-0032/2020\|EPI_ISL_540932\|202 | ···················· | ···················· | ····················· | ···················· | ···················· | ···················· |
| H | 3007 | hCoV-19/Peru/LIM-UPCH-0033/2020\|EPI_ISL_540933\|202 | ···················· | ···················· | ····················· | ···················· | ···················· | ···················· |
| H | 3008 | hCoV-19/Peru/LIM-UPCH-0034/2020\|EPI_ISL_540934\|202 | ···················· | ···················· | ····················· | ···················· | ···················· | ···················· |

| | | | | | | | |
|---|---|---|---|---|---|---|---|
| H | 3009 hCoV-19/Peru/CAL-UPCH-0035/2020\|EPI_ISL_540935\|202 | ···················· | ···················· | ····················· | ···················· | ···················· | ···················· |
| H | 3010 hCoV-19/Peru/LIM-UPCH-0045/2020\|EPI_ISL_540936\|202 | ···················· | ···················· | ····················· | ···················· | ···················· | ···················· |
| H | 3011 hCoV-19/Peru/CAL-UPCH-0046/2020\|EPI_ISL_540937\|202 | ···················· | ···················· | ····················· | ···················· | ···················· | ···················· |
| H | 3012 hCoV-19/Peru/CAL-UPCH-0047/2020\|EPI_ISL_540938\|202 | ···················· | ···················· | ····················· | ···················· | ···················· | ···················· |
| H | 3013 hCoV-19/Peru/LIM-UPCH-0050/2020\|EPI_ISL_540939\|202 | ···················· | ···················· | ····················· | ···················· | ···················· | ···················· |
| H | 3014 hCoV-19/Peru/ANC-UPCH-0051/2020\|EPI_ISL_540940\|202 | ···················· | ···················· | ····················· | ···················· | ···················· | ···················· |
| H | 3015 hCoV-19/Peru/CAL-UPCH-0053/2020\|EPI_ISL_540941\|202 | ···················· | ···················· | ····················· | ···················· | ···················· | ···················· |
| H | 3016 hCoV-19/Peru/LIM-UPCH-0056/2020\|EPI_ISL_540942\|202 | ···················· | ···················· | ····················· | ···················· | ···················· | ···················· |
| H | 3017 hCoV-19/Peru/LIM-UPCH-0057/2020\|EPI_ISL_540943\|202 | ···················· | ···················· | ····················· | ···················· | ···················· | ···················· |
| H | 3018 hCoV-19/Peru/LIM-UPCH-0059/2020\|EPI_ISL_540944\|202 | ···················· | ···················· | ····················· | ···················· | ···················· | ···················· |
| H | 3019 hCoV-19/Peru/LIM-UPCH-0061/2020\|EPI_ISL_540945\|202 | ···················· | ···················· | ····················· | ···················· | ···················· | ···················· |
| H | 3020 hCoV-19/Peru/CAL-UPCH-0062/2020\|EPI_ISL_540946\|202 | ···················· | ···················· | ····················· | ···················· | ···················· | ···················· |
| H | 3021 hCoV-19/Peru/LIM-UPCH-0063/2020\|EPI_ISL_540947\|202 | ···················· | ···················· | ····················· | ···················· | ···················· | ···················· |
| H | 3022 hCoV-19/Peru/LIM-UPCH-0065/2020\|EPI_ISL_540948\|202 | ···················· | ···················· | ····················· | ···················· | ···················· | ···················· |
| H | 3023 hCoV-19/Peru/LIM-UPCH-0066/2020\|EPI_ISL_540949\|202 | ···················· | ···················· | ····················· | ···················· | ···················· | ···················· |
| H | 3024 hCoV-19/Peru/UCA-UPCH-0067/2020\|EPI_ISL_540950\|202 | ···················· | ···················· | ····················· | ···················· | ···················· | ···················· |
| H | 3025 hCoV-19/Peru/LIM-UPCH-0068/2020\|EPI_ISL_540951\|202 | ···················· | ···················· | ····················· | ···················· | ···················· | ···················· |
| H | 3026 hCoV-19/Peru/LIM-UPCH-0070/2020\|EPI_ISL_540952\|202 | ···················· | ···················· | ····················· | ···················· | ···················· | ···················· |
| H | 3027 hCoV-19/Peru/CAL-UPCH-0071/2020\|EPI_ISL_540953\|202 | ···················· | ···················· | ····················· | ···················· | ···················· | ···················· |
| H | 3028 hCoV-19/Peru/LIM-UPCH-0073/2020\|EPI_ISL_540954\|202 | ···················· | ···················· | ····················· | ···················· | ···················· | ···················· |
| H | 3029 hCoV-19/Peru/LIM-UPCH-0074/2020\|EPI_ISL_540955\|202 | ···················· | ···················· | ····················· | ···················· | ···················· | ···················· |
| H | 3030 hCoV-19/Peru/LIM-UPCH-0078/2020\|EPI_ISL_540956\|202 | ···················· | ·A·················· | ····················· | ···················· | ···················· | ···················· |
| H | 3031 hCoV-19/Peru/LIM-UPCH-0079/2020\|EPI_ISL_540957\|202 | ···················· | ···················· | ····················· | ···················· | ···················· | ···················· |
| H | 3032 hCoV-19/Peru/CAL-UPCH-0080/2020\|EPI_ISL_540958\|202 | ···················· | ···················· | ····················· | ···················· | ···················· | ···················· |
| H | 3033 hCoV-19/Peru/LIM-UPCH-0081/2020\|EPI_ISL_540959\|202 | ···················· | ···················· | ····················· | ···················· | ···················· | ···················· |
| H | 3035 hCoV-19/USA/TX-HMH-3657/2020\|EPI_ISL_543101\|2020-0 | ···················· | ···················· | ····················· | ···················· | ···················· | ···················· |
| H | 3036 hCoV-19/USA/TX-HMH-3658/2020\|EPI_ISL_543102\|2020-0 | ···················· | ···················· | ····················· | ···················· | ···················· | ···················· |
| H | 3037 hCoV-19/USA/TX-HMH-3660/2020\|EPI_ISL_543104\|2020-0 | ···················· | ·············A······ | ····················· | ···················· | ···················· | ···················· |
| H | 3039 hCoV-19/USA/TX-HMH-3687/2020\|EPI_ISL_543107\|2020-0 | ···················· | ···················· | ····················· | ···················· | ···················· | ···················· |
| H | 3046 hCoV-19/USA/TX-HMH-3697/2020\|EPI_ISL_543116\|2020-0 | ···················· | ···················· | ····················· | ···················· | ···················· | ···················· |
| H | 3052 hCoV-19/USA/TX-HMH-3706/2020\|EPI_ISL_543123\|2020-0 | ···················· | ···················· | ····················· | ···················· | ···················· | ···················· |
| H | 3054 hCoV-19/USA/TX-HMH-3709/2020\|EPI_ISL_543126\|2020-0 | ···················· | ·············A······ | ····················· | ···················· | ···················· | ···················· |
| H | 3056 hCoV-19/USA/TX-HMH-3714/2020\|EPI_ISL_543131\|2020-0 | ···················· | ···················· | ····················· | ···················· | ···················· | ···················· |
| H | 3057 hCoV-19/USA/TX-HMH-3715/2020\|EPI_ISL_543132\|2020-0 | ···················· | ···················· | ····················· | ···················· | ···················· | ·······NN··········· |
| H | 3058 hCoV-19/USA/TX-HMH-3716/2020\|EPI_ISL_543133\|2020-0 | ···················· | ···················· | ····················· | ···················· | ···················· | ···················· |
| H | 3061 hCoV-19/USA/TX-HMH-3722/2020\|EPI_ISL_543138\|2020-0 | ···················· | ···················· | ····················· | ···················· | ···················· | ···················· |
| H | 3062 hCoV-19/USA/TX-HMH-3734/2020\|EPI_ISL_543146\|2020-0 | ···················· | ···················· | ····················· | ···················· | ···················· | ···A················ |
| H | 3063 hCoV-19/USA/TX-HMH-3735/2020\|EPI_ISL_543147\|2020-0 | ···················· | ·············A······ | ····················· | ···················· | ···················· | ···················· |
| H | 3065 hCoV-19/USA/TX-HMH-3785/2020\|EPI_ISL_543190\|2020-0 | ···················· | ···················· | ····················· | ···················· | ···················· | ···················· |
| H | 3066 hCoV-19/USA/TX-HMH-3786/2020\|EPI_ISL_543191\|2020-0 | ···················· | ···················· | ····················· | ···················· | ···················· | ···················· |
| H | 3067 hCoV-19/USA/TX-HMH-3788/2020\|EPI_ISL_543193\|2020-0 | ···················· | ···················· | ····················· | ···················· | ···················· | ···················· |
| H | 3068 hCoV-19/USA/TX-HMH-3789/2020\|EPI_ISL_543194\|2020-0 | ···················· | ···················· | ····················· | ···················· | ···················· | ···················· |
| H | 3070 hCoV-19/USA/TX-HMH-3792/2020\|EPI_ISL_543197\|2020-0 | ···················· | ···················· | ····················· | ···················· | ···················· | ···················· |
| H | 3072 hCoV-19/USA/TX-HMH-3795/2020\|EPI_ISL_543199\|2020-0 | ···················· | ···················· | ····················· | ···················· | ···················· | ···················· |
| H | 3073 hCoV-19/England/QEUH-964FF5/2020\|EPI_ISL_531161\|20 | ···················· | ···················· | ····················· | ···················· | ···················· | ···················· |
| H | 3074 hCoV-19/England/QEUH-9653CB/2020\|EPI_ISL_531163\|20 | ···················· | ···················· | ····················· | ···················· | ···················· | ···················· |
| H | 3075 hCoV-19/England/QEUH-965844/2020\|EPI_ISL_531164\|20 | ···················· | ···················· | ····················· | ···················· | ···················· | ···················· |
| H | 3076 hCoV-19/England/QEUH-964B00/2020\|EPI_ISL_531165\|20 | ···················· | ···················· | ····················· | ···················· | ···················· | ···················· |
| H | 3077 hCoV-19/Scotland/QEUH-96548C/2020\|EPI_ISL_531166\|2 | ···················· | ···················· | ····················· | ···················· | ···················· | ···················· |
| H | 3078 hCoV-19/England/QEUH-964B88/2020\|EPI_ISL_531167\|20 | ···················· | ···················· | ····················· | ···················· | ···················· | ···················· |
| H | 3079 hCoV-19/Northern Ireland/QEUH-964E07/2020\|EPI_ISL_ | ···················· | ···················· | ····················· | ···················· | ···················· | ···················· |
| H | 3080 hCoV-19/England/QEUH-96545F/2020\|EPI_ISL_531169\|20 | ···················· | ···················· | ····················· | ···················· | ···················· | ···················· |
| H | 3081 hCoV-19/England/QEUH-965237/2020\|EPI_ISL_531170\|20 | ···················· | ···················· | ····················· | ···················· | ···················· | ···················· |
| H | 3082 hCoV-19/England/QEUH-969B92/2020\|EPI_ISL_531171\|20 | ···················· | ···················· | ····················· | ···················· | ···················· | ···················· |
| H | 3083 hCoV-19/Northern Ireland/QEUH-9696FB/2020\|EPI_ISL_ | ···················· | ·A·················· | ····················· | ···················· | ···················· | ···················· |
| H | 3084 hCoV-19/Scotland/QEUH-9695C1/2020\|EPI_ISL_531174\|2 | ···················· | ···················· | ····················· | ···················· | ···················· | ···················· |
| H | 3085 hCoV-19/Scotland/QEUH-9698B9/2020\|EPI_ISL_531175\|2 | ···················· | ···················· | ····················· | ···················· | ···················· | ···················· |
| H | 3086 hCoV-19/England/QEUH-9651C1/2020\|EPI_ISL_531176\|20 | ···················· | ···················· | ····················· | ···················· | ···················· | ···················· |
| H | 3087 hCoV-19/England/QEUH-96561D/2020\|EPI_ISL_531177\|20 | ···················· | ···················· | ····················· | ···················· | ······T············· | ···················· |
| H | 3088 hCoV-19/England/QEUH-965343/2020\|EPI_ISL_531178\|20 | ···················· | ···················· | ····················· | ···················· | ···················· | ···················· |
| H | 3089 hCoV-19/England/QEUH-965440/2020\|EPI_ISL_531179\|20 | ···················· | ···················· | ····················· | ···················· | ···················· | ···················· |
| H | 3090 hCoV-19/England/QEUH-9655C5/2020\|EPI_ISL_531200\|20 | ···················· | ···················· | ····················· | ···················· | ···················· | ···················· |
| H | 3091 hCoV-19/England/QEUH-964C3A/2020\|EPI_ISL_531201\|20 | ···················· | ···················· | ····················· | ···················· | ···················· | ···················· |
| H | 3092 hCoV-19/England/QEUH-965307/2020\|EPI_ISL_531203\|20 | ···················· | ···················· | ····················· | ···················· | ···················· | ···················· |
| H | 3093 hCoV-19/England/QEUH-9653DA/2020\|EPI_ISL_531206\|20 | ···················· | ···················· | ····················· | ···················· | ···················· | ···················· |
| H | 3094 hCoV-19/Scotland/QEUH-964A4F/2020\|EPI_ISL_531207\|2 | ···················· | ···················· | ····················· | ···················· | ···················· | ···················· |
| H | 3095 hCoV-19/England/QEUH-964BA6/2020\|EPI_ISL_531209\|20 | ···················· | ·A·················· | ····················· | ···················· | ···················· | ···················· |
| H | 3096 hCoV-19/Wales/QEUH-969488/2020\|EPI_ISL_531210\|2020 | ···················· | ···················· | ····················· | ···················· | ···················· | ···················· |
| H | 3097 hCoV-19/Scotland/QEUH-969725/2020\|EPI_ISL_531212\|2 | ···················· | ·A·················· | ····················· | ···················· | ···················· | ···················· |
| H | 3098 hCoV-19/England/QEUH-964A12/2020\|EPI_ISL_531213\|20 | ···················· | ···················· | ····················· | ···················· | ···················· | ···················· |
| H | 3099 hCoV-19/England/QEUH-95F8F2/2020\|EPI_ISL_532491\|20 | ···················· | ···················· | ····················· | ···················· | ···················· | ···················· |
| H | 3100 hCoV-19/Scotland/QEUH-95FBCC/2020\|EPI_ISL_532492\|2 | ···················· | ···················· | ····················· | ···················· | ···················· | ···················· |
| H | 3101 hCoV-19/Scotland/QEUH-95E88A/2020\|EPI_ISL_532493\|2 | ···················· | ···················· | ····················· | ···················· | ···················· | ···················· |
| H | 3102 hCoV-19/England/QEUH-95F92C/2020\|EPI_ISL_532494\|20 | ···················· | ···················· | ····················· | ···················· | ···················· | ···················· |
| H | 3103 hCoV-19/England/QEUH-95F4A3/2020\|EPI_ISL_532495\|20 | ···················· | ·A·················· | ····················· | ···················· | ···················· | ···················· |
| H | 3104 hCoV-19/Northern Ireland/QEUH-95EC42/2020\|EPI_ISL_ | ···················· | ···················· | ····················· | ···················· | ···················· | ···················· |
| H | 3105 hCoV-19/England/QEUH-95F7B9/2020\|EPI_ISL_532497\|20 | ···················· | ···················· | ····················· | ···················· | ···················· | ···················· |
| H | 3106 hCoV-19/Northern Ireland/QEUH-95EB45/2020\|EPI_ISL_ | ···················· | ···················· | ····················· | ···················· | ···················· | ···················· |
| H | 3107 hCoV-19/England/QEUH-95F009/2020\|EPI_ISL_532499\|20 | ···················· | ···················· | ····················· | ···················· | ···················· | ···················· |
| H | 3108 hCoV-19/England/QEUH-95F0BE/2020\|EPI_ISL_532500\|20 | ···················· | ···················· | ····················· | ···················· | ···················· | ···················· |

| | | | | | | | | | |
|---|---|---|---|---|---|---|---|---|---|
| H | 3109 | hCoV-19/England/QEUH-95F4B2/2020\|EPI_ISL_532501\|20 | ····················· | ····················· | ····················· | ····················· | ····················· | ····················· | |
| H | 3110 | hCoV-19/Northern Ireland/QEUH-95EC06/2020\|EPI_ISL_ | ····················· | ····················· | ····················· | ····················· | ····················· | ····················· | |
| H | 3111 | hCoV-19/England/QEUH-95F458/2020\|EPI_ISL_532503\|20 | ····················· | ····················· | ····················· | ····················· | ····················· | ····················· | |
| H | 3112 | hCoV-19/England/QEUH-95F31F/2020\|EPI_ISL_532504\|20 | ····················· | ····················· | ····················· | ····················· | ····················· | ····················· | |
| H | 3113 | hCoV-19/England/QEUH-95FAED/2020\|EPI_ISL_532505\|20 | ····················· | ····················· | ····················· | ····················· | ····················· | ····················· | |
| H | 3114 | hCoV-19/Scotland/QEUH-95E714/2020\|EPI_ISL_532506\|2 | ····················· | ····················· | ····················· | ····················· | ····················· | ····················· | |
| H | 3115 | hCoV-19/England/QEUH-95E7BA/2020\|EPI_ISL_532507\|20 | ····················· | ····················· | ····················· | ····················· | ····················· | ····················· | |
| H | 3116 | hCoV-19/Northern Ireland/QEUH-95FA0B/2020\|EPI_ISL_ | ····················· | ····················· | ····················· | ····················· | ····················· | ····················· | |
| H | 3117 | hCoV-19/England/QEUH-95F801/2020\|EPI_ISL_532509\|20 | ····················· | ····················· | ····················· | ····················· | ····················· | ····················· | |
| H | 3118 | hCoV-19/Scotland/QEUH-95EA2A/2020\|EPI_ISL_532510\|2 | ····················· | ····················· | ····················· | ····················· | ····················· | ····················· | |
| H | 3119 | hCoV-19/Scotland/QEUH-95E85D/2020\|EPI_ISL_532512\|2 | ····················· | ····················· | ····················· | ····················· | ····················· | ····················· | |
| H | 3120 | hCoV-19/England/QEUH-95F124/2020\|EPI_ISL_532513\|20 | ····················· | ·A··················· | ····················· | ····················· | ····················· | ····················· | |
| H | 3121 | hCoV-19/Scotland/QEUH-95EA0C/2020\|EPI_ISL_532514\|2 | ····················· | ····················· | ····················· | ····················· | ····················· | ····················· | |
| H | 3122 | hCoV-19/Scotland/QEUH-95F4FE/2020\|EPI_ISL_532515\|2 | ····················· | ····················· | ····················· | ····················· | ····················· | ····················· | |
| H | 3123 | hCoV-19/England/QEUH-95EE3D/2020\|EPI_ISL_532516\|20 | ····················· | ····················· | ····················· | ····················· | ····················· | ····················· | |
| H | 3124 | hCoV-19/England/QEUH-95EBCD/2020\|EPI_ISL_532517\|20 | ····················· | ····················· | ····················· | ····················· | ····················· | ····················· | |
| H | 3125 | hCoV-19/Scotland/QEUH-95E662/2020\|EPI_ISL_532518\|2 | ····················· | ····················· | ····················· | ····················· | ····················· | ····················· | |
| H | 3126 | hCoV-19/Northern Ireland/QEUH-95EB81/2020\|EPI_ISL_ | ····················· | ····················· | ····················· | ····················· | ····················· | ····················· | |
| H | 3127 | hCoV-19/England/QEUH-95F072/2020\|EPI_ISL_532521\|20 | ····················· | ····················· | ····················· | ····················· | ····················· | ····················· | |
| H | 3128 | hCoV-19/England/QEUH-95EEB5/2020\|EPI_ISL_532522\|20 | ····················· | ····················· | ····················· | ····················· | ····················· | ····················· | |
| H | 3129 | hCoV-19/Scotland/QEUH-95F106/2020\|EPI_ISL_532523\|2 | ····················· | ····················· | ····················· | ····················· | ····················· | ····················· | |
| H | 3130 | hCoV-19/England/QEUH-95F42B/2020\|EPI_ISL_532524\|20 | ····················· | ····················· | ····················· | ····················· | ····················· | ····················· | |
| H | 3131 | hCoV-19/England/QEUH-95F28B/2020\|EPI_ISL_532525\|20 | ····················· | ····················· | ····················· | ····················· | ····················· | ····················· | |
| H | 3132 | hCoV-19/England/QEUH-95EE4C/2020\|EPI_ISL_532526\|20 | ····················· | ·A·······A··········· | ····················· | ····················· | ····················· | ····················· | |
| H | 3133 | hCoV-19/England/QEUH-95FB80/2020\|EPI_ISL_532527\|20 | ····················· | ····················· | ····················· | ····················· | ····················· | ····················· | |
| H | 3134 | hCoV-19/England/QEUH-95EF2B/2020\|EPI_ISL_532528\|20 | ····················· | ····················· | ····················· | ····················· | ····················· | ····················· | |
| H | 3135 | hCoV-19/England/QEUH-95E7C9/2020\|EPI_ISL_532529\|20 | ····················· | ·A··················· | ····················· | ····················· | ····················· | ····················· | |
| H | 3136 | hCoV-19/England/QEUH-95F133/2020\|EPI_ISL_532530\|20 | ····················· | ·A··················· | ····················· | ····················· | ····················· | ····················· | |
| H | 3137 | hCoV-19/Scotland/QEUH-95F4EF/2020\|EPI_ISL_532531\|2 | ····················· | ····················· | ····················· | ····················· | ····················· | ····················· | |
| H | 3138 | hCoV-19/England/QEUH-95EF58/2020\|EPI_ISL_532532\|20 | ····················· | ·A······CA··········· | ····················· | ····················· | ····················· | ····················· | 1 |
| H | 3139 | hCoV-19/England/QEUH-95F82F/2020\|EPI_ISL_532533\|20 | ····················· | ····················· | ····················· | ····················· | ····················· | ····················· | |
| H | 3140 | hCoV-19/England/QEUH-95EFEF/2020\|EPI_ISL_532534\|20 | ····················· | ····················· | ····················· | ····················· | ····················· | ····················· | |
| H | 3141 | hCoV-19/England/QEUH-95EE5B/2020\|EPI_ISL_532535\|20 | ····················· | ·A··················· | ····················· | ····················· | ····················· | ····················· | |
| H | 3142 | hCoV-19/England/QEUH-95F643/2020\|EPI_ISL_532536\|20 | ····················· | ····················· | ····················· | ····················· | ····················· | ····················· | |
| H | 3143 | hCoV-19/Northern Ireland/QEUH-95EB63/2020\|EPI_ISL_ | ····················· | ····················· | ····················· | ····················· | ····················· | ····················· | |
| H | 3144 | hCoV-19/Scotland/QEUH-95E6F9/2020\|EPI_ISL_532538\|2 | ····················· | ····················· | ····················· | ····················· | ····················· | ····················· | |
| H | 3145 | hCoV-19/England/QEUH-943A73/2020\|EPI_ISL_532539\|20 | ····················· | ····················· | ····················· | ····················· | ····················· | ····················· | |
| H | 3146 | hCoV-19/Scotland/QEUH-9439B2/2020\|EPI_ISL_532540\|2 | ····················· | ····················· | ····················· | ····················· | ····················· | ····················· | |
| H | 3147 | hCoV-19/England/QEUH-9436BB/2020\|EPI_ISL_532541\|20 | ····················· | ····················· | ····················· | ····················· | ····················· | ····················· | |
| H | 3148 | hCoV-19/Northern Ireland/QEUH-943BCB/2020\|EPI_ISL_ | ····················· | ····················· | ····················· | ····················· | ····················· | ····················· | |
| H | 3149 | hCoV-19/England/QEUH-943466/2020\|EPI_ISL_532543\|20 | ····················· | ····················· | ····················· | ····················· | ····················· | ····················· | |
| H | 3150 | hCoV-19/England/QEUH-9435BE/2020\|EPI_ISL_532544\|20 | ····················· | ····················· | ····················· | ····················· | ····················· | ····················· | |
| H | 3151 | hCoV-19/Canada/QC-LSPQ-L00232917/2020\|EPI_ISL_5360 | ····················· | ····················· | ····················· | ····················· | ····················· | ····················· | |
| H | 3152 | hCoV-19/Canada/QC-LSPQ-L00232925/2020\|EPI_ISL_5360 | ····················· | ····················· | ····················· | ····················· | ····················· | ····················· | |
| H | 3153 | hCoV-19/Canada/QC-LSPQ-L00232941/2020\|EPI_ISL_5360 | ····················· | ····················· | ····················· | ····················· | ····················· | ····················· | |
| H | 3154 | hCoV-19/Canada/QC-LSPQ-L00232949/2020\|EPI_ISL_5360 | ····················· | ····················· | ····················· | ····················· | ····················· | ····················· | |
| H | 3155 | hCoV-19/Canada/QC-LSPQ-L00232952/2020\|EPI_ISL_5360 | ····················· | ····················· | ····················· | ····················· | ····················· | ····················· | |
| H | 3156 | hCoV-19/Canada/QC-LSPQ-L00232955/2020\|EPI_ISL_5360 | ····················· | ····················· | ····················· | ····················· | ····················· | ····················· | |
| H | 3157 | hCoV-19/Canada/QC-LSPQ-L00233003/2020\|EPI_ISL_5360 | ····················· | ····················· | ····················· | ····················· | ····················· | ····················· | |
| H | 3158 | hCoV-19/Canada/QC-LSPQ-L00233038/2020\|EPI_ISL_5360 | ····················· | ····················· | ····················· | ····················· | ····················· | ····················· | |
| H | 3159 | hCoV-19/Canada/QC-LSPQ-L00233063/2020\|EPI_ISL_5360 | ····················· | ····················· | ····················· | ····················· | ····················· | ····················· | |
| H | 3160 | hCoV-19/Canada/QC-LSPQ-L00233115/2020\|EPI_ISL_5360 | ····················· | ····················· | ····················· | ····················· | ····················· | ····················· | |
| H | 3161 | hCoV-19/Canada/QC-LSPQ-L00233169/2020\|EPI_ISL_5360 | ····················· | ····················· | ····················· | ····················· | ····················· | ····················· | |
| H | 3162 | hCoV-19/Canada/QC-LSPQ-L00233171/2020\|EPI_ISL_5360 | ····················· | ····················· | ····················· | ····················· | ····················· | ····················· | |
| H | 3163 | hCoV-19/Canada/QC-LSPQ-L00233253/2020\|EPI_ISL_5360 | ····················· | ····················· | ····················· | ····················· | ····················· | ····················· | |
| H | 3164 | hCoV-19/Canada/QC-LSPQ-L00233281/2020\|EPI_ISL_5360 | ····················· | ····················· | ····················· | ····················· | ····················· | ····················· | |
| H | 3165 | hCoV-19/Canada/QC-LSPQ-L00233316/2020\|EPI_ISL_5360 | ····················· | ····················· | ····················· | ····················· | ····················· | ····················· | |
| H | 3166 | hCoV-19/Canada/QC-LSPQ-L00233323/2020\|EPI_ISL_5360 | ····················· | ····················· | ····················· | ····················· | ····················· | ····················· | |
| H | 3167 | hCoV-19/Canada/QC-LSPQ-L00233333/2020\|EPI_ISL_5360 | ····················· | ····················· | ····················· | ····················· | ····················· | ····················· | |
| H | 3168 | hCoV-19/Canada/QC-LSPQ-L00233340/2020\|EPI_ISL_5360 | ····················· | ····················· | ····················· | ····················· | ····················· | ····················· | |
| H | 3169 | hCoV-19/Canada/QC-LSPQ-L00233357/2020\|EPI_ISL_5360 | ····················· | ····················· | ····················· | ····················· | ····················· | ····················· | |
| H | 3170 | hCoV-19/Canada/QC-LSPQ-L00233380/2020\|EPI_ISL_5360 | ····················· | ····················· | ····················· | ····················· | ····················· | ····················· | |
| H | 3171 | hCoV-19/Canada/QC-LSPQ-L00233398/2020\|EPI_ISL_5360 | ····················· | ····················· | ····················· | ····················· | ····················· | ····················· | |
| H | 3172 | hCoV-19/Canada/QC-LSPQ-L00233422/2020\|EPI_ISL_5360 | ····················· | ····················· | ····················· | ····················· | ····················· | ····················· | |
| H | 3173 | hCoV-19/Canada/QC-LSPQ-L00233493/2020\|EPI_ISL_5360 | ····················· | ····················· | ····················· | ····················· | ····················· | ····················· | |
| H | 3174 | hCoV-19/Canada/QC-LSPQ-L00233496/2020\|EPI_ISL_5360 | ····················· | ····················· | ····················· | ····················· | ····················· | ····················· | |
| H | 3175 | hCoV-19/Canada/QC-LSPQ-L00233527/2020\|EPI_ISL_5360 | ····················· | ····················· | ····················· | ····················· | ····················· | ····················· | |
| H | 3176 | hCoV-19/Canada/QC-LSPQ-L00233567/2020\|EPI_ISL_5360 | ····················· | ····················· | ····················· | ····················· | ····················· | ····················· | |
| H | 3177 | hCoV-19/Canada/QC-LSPQ-L00233585/2020\|EPI_ISL_5360 | ····················· | ····················· | ····················· | ····················· | ····················· | ····················· | |
| H | 3178 | hCoV-19/Canada/QC-LSPQ-L00233633/2020\|EPI_ISL_5360 | ····················· | ····················· | ····················· | ····················· | ····················· | ····················· | |
| H | 3179 | hCoV-19/Canada/QC-LSPQ-L00233661/2020\|EPI_ISL_5360 | ····················· | ·A··················· | ····················· | ····················· | ····················· | ····················· | |
| H | 3180 | hCoV-19/Canada/QC-LSPQ-L00233681/2020\|EPI_ISL_5360 | ····················· | ····················· | ····················· | ····················· | ····················· | ····················· | |
| H | 3181 | hCoV-19/Canada/QC-LSPQ-L00233683/2020\|EPI_ISL_5360 | ····················· | ····················· | ····················· | ····················· | ····················· | ····················· | |
| H | 3182 | hCoV-19/Canada/QC-LSPQ-L00233695/2020\|EPI_ISL_5360 | ····················· | ·A··················· | ····················· | ····················· | ····················· | ····················· | |
| H | 3183 | hCoV-19/Canada/QC-LSPQ-L00233702/2020\|EPI_ISL_5360 | ····················· | ····················· | ····················· | ····················· | ····················· | ····················· | |
| H | 3184 | hCoV-19/Canada/QC-LSPQ-L00233714/2020\|EPI_ISL_5360 | ····················· | ····················· | ····················· | ····················· | ····················· | ····················· | |
| H | 3185 | hCoV-19/Canada/QC-LSPQ-L00233721/2020\|EPI_ISL_5360 | ····················· | ····················· | ····················· | ····················· | ····················· | ····················· | |
| H | 3186 | hCoV-19/Canada/QC-LSPQ-L00233734/2020\|EPI_ISL_5360 | ····················· | ····················· | ····················· | ····················· | ····················· | ····················· | |
| H | 3187 | hCoV-19/Canada/QC-LSPQ-L00233793/2020\|EPI_ISL_5360 | ····················· | ····················· | ····················· | ····················· | ····················· | ····················· | |
| H | 3188 | hCoV-19/Canada/QC-LSPQ-L00233835/2020\|EPI_ISL_5360 | ····················· | ····················· | ····················· | ····················· | ····················· | ····················· | |

| | | | | | | | | |
|---|---|---|---|---|---|---|---|---|
| H | 3189 | hCoV-19/Canada/QC-LSPQ-L00234790/2020\|EPI_ISL_5360 | .................... | .................... | .................... | .................... | .................... | .................... |
| H | 3190 | hCoV-19/Canada/QC-LSPQ-L00234851/2020\|EPI_ISL_5360 | .................... | .................... | .................... | .................... | .................... | .................... |
| H | 3191 | hCoV-19/Canada/QC-LSPQ-L00234853/2020\|EPI_ISL_5360 | .................... | .................... | .................... | .................... | .................... | .................... |
| H | 3192 | hCoV-19/Canada/QC-LSPQ-L00234865/2020\|EPI_ISL_5360 | .................... | .................... | .................... | .................... | .................... | .................... |
| H | 3194 | hCoV-19/Canada/QC-LSPQ-L00234872/2020\|EPI_ISL_5360 | .................... | .................... | .................... | .................... | .................... | .................... |
| H | 3195 | hCoV-19/Canada/QC-LSPQ-L00234877/2020\|EPI_ISL_5360 | .................... | .................... | .................... | .................... | .................... | .................... |
| H | 3196 | hCoV-19/Canada/QC-LSPQ-L00234878/2020\|EPI_ISL_5360 | .................... | .................... | .................... | .................... | .................... | .................... |
| H | 3197 | hCoV-19/Canada/QC-LSPQ-L00234887/2020\|EPI_ISL_5360 | .................... | .................... | .................... | .................... | .................... | .................... |
| H | 3198 | hCoV-19/Canada/QC-LSPQ-L00234905/2020\|EPI_ISL_5360 | .................... | .................... | .................... | .................... | .................... | .................... |
| H | 3199 | hCoV-19/Canada/QC-LSPQ-L00234990/2020\|EPI_ISL_5360 | .................... | .................... | .................... | .................... | .................... | .................... |
| H | 3200 | hCoV-19/Canada/QC-LSPQ-L00235020/2020\|EPI_ISL_5360 | .................... | .................... | .................... | .................... | .................... | .................... |
| H | 3201 | hCoV-19/Canada/QC-LSPQ-L00235027/2020\|EPI_ISL_5360 | .................... | .................... | .................... | .................... | .................... | .................... |
| H | 3202 | hCoV-19/Canada/QC-LSPQ-L00235107/2020\|EPI_ISL_5360 | .................... | .................... | .................... | .................... | .................... | .................... |
| H | 3203 | hCoV-19/Canada/QC-LSPQ-L00235132/2020\|EPI_ISL_5360 | .................... | .................... | .................... | .................... | .................... | .................... |
| H | 3204 | hCoV-19/Canada/QC-LSPQ-L00235136/2020\|EPI_ISL_5360 | .................... | .................... | .................... | .................... | .................... | .................... |
| H | 3205 | hCoV-19/Canada/QC-LSPQ-L00235218/2020\|EPI_ISL_5360 | .................... | .................... | .................... | .................... | .................... | .................... |
| H | 3206 | hCoV-19/Canada/QC-LSPQ-L00235261/2020\|EPI_ISL_5360 | .................... | .................... | .................... | .................... | .................... | .................... |
| H | 3207 | hCoV-19/Canada/QC-LSPQ-L00235295/2020\|EPI_ISL_5360 | .................... | .................... | .................... | .................... | .................... | .................... |
| H | 3208 | hCoV-19/Canada/QC-LSPQ-L00235378/2020\|EPI_ISL_5360 | .................... | .................... | .................... | .................... | .................... | .................... |
| H | 3209 | hCoV-19/Canada/QC-LSPQ-L00235380/2020\|EPI_ISL_5360 | .................... | .................... | .................... | .................... | .................... | .................... |
| H | 3210 | hCoV-19/Canada/QC-LSPQ-L00235424/2020\|EPI_ISL_5360 | .................... | .................... | .................... | .................... | .................... | .................... |
| H | 3211 | hCoV-19/Canada/QC-LSPQ-L00235430/2020\|EPI_ISL_5360 | .................... | .................... | .................... | .................... | .................... | .................... |
| H | 3212 | hCoV-19/Canada/QC-LSPQ-L00235552/2020\|EPI_ISL_5360 | .................... | .................... | .................... | .................... | .................... | .................... |
| H | 3250 | hCoV-19/Spain/CL-IBV-97006218/2020\|EPI_ISL_537364\| | .................... | .................... | .................... | .................... | .................... | .................... |
| H | 3257 | hCoV-19/Scotland/QEUH-98FA99/2020\|EPI_ISL_539980\|2 | .................... | .................... | .................... | .................... | .................... | .................... |
| H | 3258 | hCoV-19/England/QEUH-9982B3/2020\|EPI_ISL_539981\|20 | .................... | .................... | .................... | .................... | .................... | .................... |
| H | 3259 | hCoV-19/England/QEUH-990154/2020\|EPI_ISL_539983\|20 | .................... | .................... | .................... | .................... | .................... | .................... |
| H | 3260 | hCoV-19/England/QEUH-998462/2020\|EPI_ISL_539984\|20 | .................... | .................... | .................... | .................... | .................... | .................... |
| H | 3261 | hCoV-19/England/QEUH-99871E/2020\|EPI_ISL_539985\|20 | .................... | .................... | .................... | .................... | .................... | .................... |
| H | 3262 | hCoV-19/England/QEUH-998C5B/2020\|EPI_ISL_539986\|20 | .................... | .................... | .................... | .................... | .................... | .................... |
| H | 3263 | hCoV-19/England/QEUH-998338/2020\|EPI_ISL_539987\|20 | .................... | .................... | .................... | .................... | .................... | .................... |
| H | 3264 | hCoV-19/England/QEUH-98F677/2020\|EPI_ISL_539988\|20 | .................... | .A.................. | .................... | .................... | .................... | .................... |
| H | 3265 | hCoV-19/England/QEUH-990075/2020\|EPI_ISL_539990\|20 | .................... | .................... | .................... | .................... | .................... | .................... |
| H | 3266 | hCoV-19/Scotland/QEUH-98FAC6/2020\|EPI_ISL_539991\|2 | .................... | .................... | .................... | .................... | .................... | .................... |
| H | 3267 | hCoV-19/England/QEUH-998550/2020\|EPI_ISL_539992\|20 | .................... | .A.................. | .................... | .................... | .................... | .................... |
| H | 3268 | hCoV-19/England/QEUH-9903E5/2020\|EPI_ISL_539993\|20 | .................... | .................... | .................... | .................... | .................... | .................... |
| H | 3269 | hCoV-19/England/QEUH-98FCC0/2020\|EPI_ISL_539994\|20 | .................... | .................... | .................... | .................... | .................... | .................... |
| H | 3270 | hCoV-19/England/QEUH-998945/2020\|EPI_ISL_539995\|20 | .................... | .A.................. | .................... | .................... | .................... | ..A................. |
| H | 3271 | hCoV-19/England/QEUH-99036D/2020\|EPI_ISL_539996\|20 | .................... | .................... | .................... | .................... | .................... | .................... |
| H | 3272 | hCoV-19/England/QEUH-9903D6/2020\|EPI_ISL_539997\|20 | .................... | .................... | .................... | .................... | .................... | .................... |
| H | 3273 | hCoV-19/England/QEUH-9903A9/2020\|EPI_ISL_539998\|20 | .................... | .................... | .................... | .................... | .................... | .................... |
| H | 3274 | hCoV-19/England/QEUH-99831A/2020\|EPI_ISL_539999\|20 | .................... | .................... | .................... | .................... | .................... | .................... |
| H | 3275 | hCoV-19/Peru/LIM-UPCH-0082/2020\|EPI_ISL_540960\|202 | .................... | .................... | .................... | .................... | .................... | .................... |
| H | 3276 | hCoV-19/Peru/LIM-UPCH-0083/2020\|EPI_ISL_540961\|202 | .................... | .................... | .................... | .................... | .................... | .................... |
| H | 3277 | hCoV-19/Peru/LIM-UPCH-0084/2020\|EPI_ISL_540962\|202 | .................... | .................... | .................... | .................... | .................... | .................... |
| H | 3278 | hCoV-19/Peru/LIM-UPCH-0085/2020\|EPI_ISL_540963\|202 | .................... | .................... | .................... | .................... | .................... | .................... |
| H | 3279 | hCoV-19/Peru/LIM-UPCH-0086/2020\|EPI_ISL_540964\|202 | .................... | .................... | .................... | .................... | .................... | .................... |
| H | 3280 | hCoV-19/Peru/LIM-UPCH-0087/2020\|EPI_ISL_540965\|202 | .................... | .................... | .................... | .................... | .................... | .................... |
| H | 3281 | hCoV-19/Peru/LIM-UPCH-0088/2020\|EPI_ISL_540966\|202 | .................... | .................... | .................... | .................... | .................... | .................... |
| H | 3282 | hCoV-19/Peru/CAL-UPCH-0089/2020\|EPI_ISL_540967\|202 | .................... | .................... | .................... | .................... | .................... | .................... |
| H | 3283 | hCoV-19/Peru/LIM-UPCH-0090/2020\|EPI_ISL_540968\|202 | .................... | .................... | .................... | .................... | .................... | .................... |
| H | 3284 | hCoV-19/Peru/LIM-UPCH-0091/2020\|EPI_ISL_540969\|202 | .................... | .................... | .................... | .................... | .................... | .................... |
| H | 3285 | hCoV-19/Peru/CAL-UPCH-0092/2020\|EPI_ISL_540970\|202 | .................... | .................... | .................... | .................... | .................... | .................... |
| H | 3286 | hCoV-19/Peru/LIM-UPCH-0093/2020\|EPI_ISL_540971\|202 | .................... | .................... | .................... | .................... | .................... | .................... |
| H | 3287 | hCoV-19/Peru/LIM-UPCH-0094/2020\|EPI_ISL_540972\|202 | .................... | .................... | .................... | .................... | .................... | .................... |
| H | 3288 | hCoV-19/Peru/LIM-UPCH-0095/2020\|EPI_ISL_540973\|202 | .................... | .................... | .................... | .................... | .................... | .................... |
| H | 3289 | hCoV-19/Peru/LIM-UPCH-0096/2020\|EPI_ISL_540974\|202 | .................... | .................... | .................... | .................... | .................... | .................... |
| H | 3290 | hCoV-19/Peru/CAL-UPCH-0097/2020\|EPI_ISL_540975\|202 | .................... | .................... | .................... | .................... | .................... | .................... |
| H | 3291 | hCoV-19/Peru/CAL-UPCH-0098/2020\|EPI_ISL_540976\|202 | .................... | .................... | .................... | .................... | .................... | .................... |
| H | 3292 | hCoV-19/Peru/LIM-UPCH-0099/2020\|EPI_ISL_540977\|202 | .................... | .................... | .................... | .................... | .................... | .................... |
| H | 3293 | hCoV-19/Peru/LIM-UPCH-0100/2020\|EPI_ISL_540978\|202 | .................... | .................... | .................... | .................... | .................... | .................... |
| H | 3294 | hCoV-19/Peru/LIM-UPCH-0101/2020\|EPI_ISL_540979\|202 | .................... | .................... | .................... | .................... | .................... | .................... |
| H | 3295 | hCoV-19/Peru/LIM-UPCH-0102/2020\|EPI_ISL_540980\|202 | .................... | .................... | .................... | .................... | .................... | .................... |
| H | 3296 | hCoV-19/Peru/LIM-UPCH-0103/2020\|EPI_ISL_540981\|202 | .................... | .................... | .................... | .................... | .................... | .................... |
| H | 3297 | hCoV-19/Peru/LIM-UPCH-0104/2020\|EPI_ISL_540982\|202 | .................... | .................... | .................... | .................... | .................... | .................... |
| H | 3298 | hCoV-19/Peru/ANC-UPCH-0105/2020\|EPI_ISL_540983\|202 | .................... | .................... | .................... | .................... | .................... | .................... |
| H | 3299 | hCoV-19/Peru/ANC-UPCH-0106/2020\|EPI_ISL_540984\|202 | .................... | .................... | .................... | .................... | .................... | .................... |
| H | 3300 | hCoV-19/Peru/ANC-UPCH-0107/2020\|EPI_ISL_540985\|202 | .................... | .................... | .................... | .................... | .................... | .................... |
| H | 3301 | hCoV-19/Peru/LAL-UPCH-0109/2020\|EPI_ISL_540986\|202 | .................... | .................... | .................... | .................... | .................... | .................... |
| H | 3302 | hCoV-19/Peru/ARE-UPCH-0110/2020\|EPI_ISL_540987\|202 | .................... | .................... | .................... | .................... | .................... | .................... |
| H | 3303 | hCoV-19/Peru/ARE-UPCH-0111/2020\|EPI_ISL_540988\|202 | .................... | .................... | .................... | .................... | .................... | .................... |
| H | 3304 | hCoV-19/Peru/LIM-UPCH-0112/2020\|EPI_ISL_540989\|202 | .................... | .................... | .................... | .................... | .................... | .................... |
| H | 3305 | hCoV-19/Peru/LIM-UPCH-0113/2020\|EPI_ISL_540990\|202 | .................... | .................... | .................... | .................... | .................... | .................... |
| H | 3306 | hCoV-19/Peru/LIM-UPCH-0114/2020\|EPI_ISL_540991\|202 | .................... | .................... | .................... | .................... | .................... | .................... |
| H | 3307 | hCoV-19/Peru/LIM-UPCH-0115/2020\|EPI_ISL_540992\|202 | .................... | .................... | .................... | .................... | .................... | .................... |
| H | 3308 | hCoV-19/South Korea/IHC597/2020\|EPI_ISL_540993\|202 | .................... | .................... | .................... | .................... | .................... | .................... |
| H | 3309 | hCoV-19/South Korea/IHC610/2020\|EPI_ISL_540994\|202 | .................... | .................... | .................... | .................... | .................... | .................... |
| H | 3310 | hCoV-19/South Korea/IHC614/2020\|EPI_ISL_540995\|202 | .................... | .................... | .................... | .................... | .................... | .................... |
| H | 3311 | hCoV-19/South Korea/IHC685/2020\|EPI_ISL_540996\|202 | .................... | .................... | .................... | .................... | .................... | .................... |
| H | 3312 | hCoV-19/South Korea/IHC719/2020\|EPI_ISL_540997\|202 | .................... | .................... | .................... | .................... | .................... | .................... |

```
H    3313 hCoV-19/South Korea/IHC1215/2020|EPI_ISL_540998|20    ····················  ····················  ····················  ····················  ····················  ····················
H    3314 hCoV-19/South Korea/IHC1217/2020|EPI_ISL_540999|20    ····················  ····················  ····················  ····················  ····················  ····················
H    3317 hCoV-19/USA/TX-HMH-3730/2020|EPI_ISL_543142|2020-0    ····················  ····················  ····················  ····················  ····················  ····················
H    3319 hCoV-19/USA/TX-HMH-3732/2020|EPI_ISL_543144|2020-0    ····················  ····················  ····················  ····················  ····················  ······NN············
H    3320 hCoV-19/USA/TX-HMH-3733/2020|EPI_ISL_543145|2020-0    ····················  ····················  ····················  ····················  ····················  ····················
H    3321 hCoV-19/USA/TX-HMH-3737/2020|EPI_ISL_543149|2020-0    ····················  ····················  ····················  ····················  ····················  ····················
H    3323 hCoV-19/USA/TX-HMH-3742/2020|EPI_ISL_543154|2020-0    ····················  ····················  ····················  ····················  ····················  ····················
H    3326 hCoV-19/USA/TX-HMH-3745/2020|EPI_ISL_543157|2020-0    ····················  ····················  ····················  ····················  ··············N·····  N···················
H    3328 hCoV-19/USA/TX-HMH-3747/2020|EPI_ISL_543159|2020-0    ····················  ····················  ····················  ····················  ····················  ····················
H    3329 hCoV-19/England/QEUH-964B2E/2020|EPI_ISL_531060|20    ····················  ····················  ····················  ····················  ····················  ····················
H    3330 hCoV-19/England/QEUH-964C85/2020|EPI_ISL_531061|20    ····················  ····················  ····················  ····················  ····················  ····················
H    3331 hCoV-19/England/QEUH-9656A4/2020|EPI_ISL_531062|20    ····················  ····················  ····················  ····················  ····················  ····················
H    3332 hCoV-19/Northern Ireland/QEUH-96996B/2020|EPI_ISL_    ····················  ····················  ····················  ····················  ····················  ····················
H    3333 hCoV-19/England/QEUH-964BF1/2020|EPI_ISL_531064|20    ····················  ····················  ····················  ····················  ····················  ····················
H    3334 hCoV-19/Scotland/QEUH-969A2C/2020|EPI_ISL_531065|2    ····················  ····················  ····················  ····················  ····················  ····················
H    3335 hCoV-19/England/QEUH-965853/2020|EPI_ISL_531067|20    ····················  ····················  ····················  ····················  ····················  ····················
H    3336 hCoV-19/England/QEUH-964AA9/2020|EPI_ISL_531068|20    ····················  ····················  ····················  ····················  ····················  ····················
H    3337 hCoV-19/England/QEUH-965316/2020|EPI_ISL_531069|20    ····················  ·A··················  ····················  ····················  ····················  ····················
H    3338 hCoV-19/England/QEUH-969497/2020|EPI_ISL_531070|20    ····················  ····················  ····················  ····················  ····················  ····················
H    3339 hCoV-19/England/QEUH-965862/2020|EPI_ISL_531071|20    ····················  ·A··················  ····················  ····················  ····················  ··A·················
H    3340 hCoV-19/Northern Ireland/QEUH-965097/2020|EPI_ISL_    ····················  ····················  ····················  ····················  ····················  ····················
H    3341 hCoV-19/England/QEUH-9695D0/2020|EPI_ISL_531073|20    ····················  ····················  ····················  ····················  ····················  ····················
H    3342 hCoV-19/England/QEUH-964B97/2020|EPI_ISL_531074|20    ····················  ·A··················  ····················  ····················  ····················  ····················
H    3343 hCoV-19/England/QEUH-965914/2020|EPI_ISL_531075|20    ····················  ·A··················  ····················  ····················  ····················  ··A·················
H    3344 hCoV-19/England/QEUH-969B47/2020|EPI_ISL_531076|20    ····················  ····················  ····················  ····················  ····················  ····················
H    3345 hCoV-19/England/QEUH-964942/2020|EPI_ISL_531077|20    ····················  ·A··················  ····················  ····················  ····················  ····················
H    3346 hCoV-19/England/QEUH-969558/2020|EPI_ISL_531078|20    ····················  ····················  ····················  ····················  ····················  ····················
H    3347 hCoV-19/Scotland/QEUH-964D46/2020|EPI_ISL_531079|2    ····················  ····················  ····················  ····················  ····················  ····················
H    3348 hCoV-19/England/QEUH-965282/2020|EPI_ISL_531080|20    ····················  ····················  ····················  ····················  ····················  ····················
H    3349 hCoV-19/England/QEUH-96589F/2020|EPI_ISL_531081|20    ····················  ····················  ····················  ····················  ····················  ····················
H    3350 hCoV-19/England/QEUH-969585/2020|EPI_ISL_531082|20    ····················  ····················  ····················  ····················  ····················  ····················
H    3351 hCoV-19/England/QEUH-965255/2020|EPI_ISL_531084|20    ····················  ····················  ····················  ····················  ····················  ····················
H    3352 hCoV-19/England/QEUH-9697CB/2020|EPI_ISL_531085|20    ····················  ····················  ····················  ····················  ····················  ····················
H    3353 hCoV-19/England/QEUH-965932/2020|EPI_ISL_531086|20    ····················  ····················  ····················  ····················  ····················  ····················
H    3354 hCoV-19/Scotland/QEUH-969752/2020|EPI_ISL_531087|2    ····················  ····················  ····················  ····················  ····················  ····················
H    3355 hCoV-19/Northern Ireland/QEUH-96988C/2020|EPI_ISL_    ····················  ····················  ····················  ····················  ····················  ····················
H    3356 hCoV-19/Scotland/QEUH-969549/2020|EPI_ISL_531091|2    ····················  ····················  ····················  ····················  ····················  ····················
H    3357 hCoV-19/England/QEUH-96555C/2020|EPI_ISL_531094|20    ····················  ····················  ····················  ····················  ····················  ····················
H    3358 hCoV-19/Northern Ireland/QEUH-969804/2020|EPI_ISL_    ····················  ····················  ····················  ····················  ····················  ····················
H    3359 hCoV-19/Scotland/QEUH-964B1F/2020|EPI_ISL_531096|2    ····················  ····················  ····················  ····················  ····················  ····················
H    3360 hCoV-19/England/QEUH-96564A/2020|EPI_ISL_531097|20    ····················  ····················  ····················  ····················  ····················  ····················
H    3361 hCoV-19/Northern Ireland/QEUH-964E34/2020|EPI_ISL_    ····················  ····················  ····················  ····················  ····················  ····················
H    3362 hCoV-19/Scotland/QEUH-96498E/2020|EPI_ISL_531099|2    ····················  ····················  ····················  ····················  ····················  ····················
H    3363 hCoV-19/Northern Ireland/QEUH-95EA93/2020|EPI_ISL_    ····················  ····················  ····················  ····················  ····················  ····················
H    3364 hCoV-19/Northern Ireland/QEUH-95EEF1/2020|EPI_ISL_    ····················  ····················  ····················  ····················  ····················  ····················
H    3365 hCoV-19/England/QEUH-95EC33/2020|EPI_ISL_532308|20    ····················  ····················  ····················  ····················  ····················  ····················
H    3366 hCoV-19/England/QEUH-95F555/2020|EPI_ISL_532309|20    ····················  ····················  ····················  ····················  ····················  ····················
H    3367 hCoV-19/England/QEUH-95F528/2020|EPI_ISL_532310|20    ····················  ····················  ····················  ····················  ····················  ····················
H    3368 hCoV-19/Scotland/QEUH-95E8F3/2020|EPI_ISL_532311|2    ····················  ····················  ····················  ····················  ····················  ····················
H    3369 hCoV-19/England/QEUH-95F537/2020|EPI_ISL_532312|20    ····················  ····················  ····················  ····················  ····················  ····················
H    3370 hCoV-19/England/QEUH-95F5BF/2020|EPI_ISL_532313|20    ····················  ····················  ····················  ····················  ····················  ····················
H    3371 hCoV-19/England/QEUH-95F93B/2020|EPI_ISL_532314|20    ····················  ····················  ····················  ····················  ····················  ····················
H    3372 hCoV-19/Scotland/QEUH-95F9B3/2020|EPI_ISL_532316|2    ····················  ····················  ····················  ····················  ····················  ····················
H    3373 hCoV-19/Scotland/QEUH-95E86C/2020|EPI_ISL_532317|2    ····················  ····················  ····················  ····················  ····················  ····················
H    3374 hCoV-19/England/QEUH-95E92D/2020|EPI_ISL_532318|20    ····················  ····················  ····················  ····················  ····················  ····················
H    3375 hCoV-19/England/QEUH-95E7D8/2020|EPI_ISL_532319|20    ····················  ····················  ····················  ····················  ····················  ····················
H    3376 hCoV-19/Northern Ireland/QEUH-95EAB1/2020|EPI_ISL_    ····················  ····················  ····················  ····················  ····················  ····················
H    3377 hCoV-19/Scotland/QEUH-95FA47/2020|EPI_ISL_532390|2    ····················  ····················  ····················  ····················  ····················  ····················
H    3378 hCoV-19/England/QEUH-95E802/2020|EPI_ISL_532391|20    ····················  ····················  ····················  ····················  ····················  ····················
H    3379 hCoV-19/Northern Ireland/QEUH-95EDC7/2020|EPI_ISL_    ····················  ····················  ····················  ····················  ····················  ····················
H    3380 hCoV-19/England/QEUH-95F19D/2020|EPI_ISL_532393|20    ····················  ····················  ····················  ····················  ····················  ····················
H    3381 hCoV-19/Scotland/QEUH-95E996/2020|EPI_ISL_532394|2    ····················  ····················  ····················  ····················  ····················  ····················
H    3382 hCoV-19/England/QEUH-95F77D/2020|EPI_ISL_532395|20    ····················  ····················  ····················  ····················  ····················  ····················
H    3383 hCoV-19/England/QEUH-95F6DA/2020|EPI_ISL_532396|20    ····················  ····················  ····················  ····················  ····················  ····················
H    3384 hCoV-19/Northern Ireland/QEUH-95E9D2/2020|EPI_ISL_    ····················  ····················  ····················  ····················  ····················  ····················
H    3385 hCoV-19/England/QEUH-95F25E/2020|EPI_ISL_532398|20    ····················  ····················  ····················  ····················  ····················  ····················
H    3386 hCoV-19/Scotland/QEUH-95F90E/2020|EPI_ISL_532399|2    ····················  ····················  ····················  ····················  ····················  ····················
H    3387 hCoV-19/England/OXON-B46BE/2020|EPI_ISL_534966|202    ····················  ····················  ····················  ····················  ····················  ····················
H    3388 hCoV-19/England/OXON-B46FA/2020|EPI_ISL_534967|202    ····················  ····················  ····················  ····················  ····················  ····················
H    3389 hCoV-19/England/OXON-B4715/2020|EPI_ISL_534969|202    ····················  ····················  ····················  ····················  ····················  ····················
H    3390 hCoV-19/England/OXON-B4724/2020|EPI_ISL_534970|202    ····················  ····················  ····················  ····················  ····················  ····················
H    3391 hCoV-19/England/OXON-B4760/2020|EPI_ISL_534971|202    ····················  ····················  ····················  ····················  ····················  ····················
H    3392 hCoV-19/England/OXON-B478E/2020|EPI_ISL_534972|202    ····················  ····················  ····················  ····················  ····················  ····················
H    3393 hCoV-19/England/OXON-B47CA/2020|EPI_ISL_534973|202    ····················  ····················  ····················  ····················  ····················  ····················
H    3394 hCoV-19/England/OXON-B47F7/2020|EPI_ISL_534975|202    ····················  ····················  ····················  ····················  ····················  ····················
H    3395 hCoV-19/England/OXON-B4803/2020|EPI_ISL_534976|202    ····················  ····················  ····················  ····················  ····················  ····················
H    3396 hCoV-19/England/OXON-B4821/2020|EPI_ISL_534978|202    ····················  ····················  ····················  ····················  ····················  ····················
H    3397 hCoV-19/England/OXON-B48A9/2020|EPI_ISL_534980|202    ····················  ····················  ····················  ····················  ····················  ····················
H    3398 hCoV-19/England/OXON-B48C7/2020|EPI_ISL_534981|202    ····················  ·············A······  ····················  ····················  ····················  ····················
H    3399 hCoV-19/England/OXON-B491F/2020|EPI_ISL_534982|202    ····················  ····················  ····················  ····················  ····················  ····················
```

| | | | | | | | | |
|---|---|---|---|---|---|---|---|---|
| H | 3400 hCoV-19/England/OXON-B49D3/2020\|EPI_ISL_534985\|202 | ···················· | ···················· | ···················· | ···················· | ···················· | ···················· | |
| H | 3401 hCoV-19/Canada/QC-LSPQ-L00233812/2020\|EPI_ISL_5360 | ···················· | ···················· | ···················· | ···················· | ···················· | ···················· | |
| H | 3402 hCoV-19/Canada/QC-LSPQ-L00233849/2020\|EPI_ISL_5360 | ···················· | ···················· | ···················· | ···················· | ···················· | ···················· | |
| H | 3403 hCoV-19/Canada/QC-LSPQ-L00233867/2020\|EPI_ISL_5360 | ···················· | ···················· | ···················· | ···················· | ···················· | ···················· | |
| H | 3404 hCoV-19/Canada/QC-LSPQ-L00233870/2020\|EPI_ISL_5360 | ···················· | ···················· | ···················· | ···················· | ···················· | ···················· | |
| H | 3405 hCoV-19/Canada/QC-LSPQ-L00233970/2020\|EPI_ISL_5360 | ···················· | ···················· | ···················· | ···················· | ···················· | ···················· | |
| H | 3406 hCoV-19/Canada/QC-LSPQ-L00234248/2020\|EPI_ISL_5360 | ···················· | ···················· | ···················· | ···················· | ···················· | ···················· | |
| H | 3407 hCoV-19/Canada/QC-LSPQ-L00234287/2020\|EPI_ISL_5360 | ···················· | ···················· | ···················· | ···················· | ···················· | ···················· | |
| H | 3408 hCoV-19/Canada/QC-LSPQ-L00234302/2020\|EPI_ISL_5360 | ···················· | ···················· | ···················· | ···················· | ···················· | ···················· | |
| H | 3409 hCoV-19/Canada/QC-LSPQ-L00234333/2020\|EPI_ISL_5360 | ···················· | ···················· | ···················· | ···················· | ···················· | ···················· | |
| H | 3410 hCoV-19/Canada/QC-LSPQ-L00234347/2020\|EPI_ISL_5360 | ···················· | ···················· | ···················· | ···················· | ···················· | ···················· | |
| H | 3412 hCoV-19/Canada/QC-LSPQ-L00234442/2020\|EPI_ISL_5360 | ···················· | ···················· | ···················· | ···················· | ···················· | ···················· | |
| H | 3413 hCoV-19/Canada/QC-LSPQ-L00234460/2020\|EPI_ISL_5360 | ···················· | ···················· | ···················· | ···················· | ···················· | ···················· | |
| H | 3414 hCoV-19/Canada/QC-LSPQ-L00234533/2020\|EPI_ISL_5360 | ···················· | ···················· | ···················· | ···················· | ···················· | ···················· | |
| H | 3415 hCoV-19/Canada/QC-LSPQ-L00234537/2020\|EPI_ISL_5360 | ···················· | ···················· | ···················· | ···················· | ···················· | ···················· | |
| H | 3416 hCoV-19/Canada/QC-LSPQ-L00234540/2020\|EPI_ISL_5360 | ···················· | ···················· | ···················· | ···················· | ···················· | ···················· | |
| H | 3417 hCoV-19/Canada/QC-LSPQ-L00234574/2020\|EPI_ISL_5360 | ···················· | ···················· | ···················· | ···················· | ···················· | ···················· | |
| H | 3418 hCoV-19/Canada/QC-LSPQ-L00234577/2020\|EPI_ISL_5360 | ···················· | ···················· | ···················· | ···················· | ···················· | ···················· | |
| H | 3419 hCoV-19/Canada/QC-LSPQ-L00234602/2020\|EPI_ISL_5360 | ···················· | ···················· | ···················· | ···················· | ···················· | ···················· | |
| H | 3420 hCoV-19/Canada/QC-LSPQ-L00234611/2020\|EPI_ISL_5360 | ···················· | ···················· | ···················· | ···················· | ···················· | ···················· | |
| H | 3421 hCoV-19/Canada/QC-LSPQ-L00234621/2020\|EPI_ISL_5360 | ···················· | ···················· | ···················· | ···················· | ···················· | ···················· | |
| H | 3422 hCoV-19/Canada/QC-LSPQ-L00234627/2020\|EPI_ISL_5360 | ···················· | ···················· | ···················· | ···················· | ···················· | ···················· | |
| H | 3423 hCoV-19/Canada/QC-LSPQ-L00234639/2020\|EPI_ISL_5360 | ···················· | ···················· | ···················· | ···················· | ···················· | ···················· | |
| H | 3424 hCoV-19/Canada/QC-LSPQ-L00234652/2020\|EPI_ISL_5360 | ···················· | ···················· | ···················· | ···················· | ···················· | ···················· | |
| H | 3425 hCoV-19/Canada/QC-LSPQ-L00234655/2020\|EPI_ISL_5360 | ···················· | ···················· | ···················· | ···················· | ···················· | ···················· | |
| H | 3426 hCoV-19/Canada/QC-LSPQ-L00234676/2020\|EPI_ISL_5360 | ···················· | ···················· | ···················· | ···················· | ···················· | ···················· | |
| H | 3427 hCoV-19/Canada/QC-LSPQ-L00234776/2020\|EPI_ISL_5360 | ···················· | ···················· | ···················· | ···················· | ···················· | ···················· | |
| H | 3431 hCoV-19/Spain/CL-IBV-97006228/2020\|EPI_ISL_537374\| | ···················· | ···················· | ···················· | ···················· | ·NNN················ | ···················· | 1 |
| H | 3433 hCoV-19/Spain/GA-IBV-97006346/2020\|EPI_ISL_537380\| | ···················· | ···················· | ···················· | ···················· | ···················· | ···················· | |
| H | 3435 hCoV-19/Spain/RI-IBV-99005837/2020\|EPI_ISL_537382\| | ···················· | ···················· | ···················· | ···················· | ···················· | ···················· | |
| H | 3437 hCoV-19/Spain/RI-IBV-99005842/2020\|EPI_ISL_537385\| | ···················· | ···················· | ···················· | ···················· | ···················· | ···················· | |
| H | 3438 hCoV-19/Spain/RI-IBV-99005844/2020\|EPI_ISL_537387\| | ···················· | ···················· | ···················· | ···················· | ···················· | ···················· | |
| H | 3439 hCoV-19/Spain/RI-IBV-99005847/2020\|EPI_ISL_537390\| | ···················· | ···················· | ···················· | ···················· | ···················· | ···················· | |
| H | 3445 hCoV-19/Indonesia/SU-NIHRD-PME4745/2020\|EPI_ISL_53 | ···················· | ···················· | ···················· | ···················· | ···················· | ··A················· | |
| H | 3446 hCoV-19/Indonesia/SU-NIHRD-PME4760/2020\|EPI_ISL_53 | ···················· | ···················· | ···················· | ···················· | ···················· | ··A················· | |
| H | 3447 hCoV-19/Indonesia/KT-NIHRD-PME4206/2020\|EPI_ISL_53 | ···················· | ···················· | ···················· | ···················· | ···················· | ···················· | |
| H | 3448 hCoV-19/Indonesia/BA-NIHRD-PME3926/2020\|EPI_ISL_53 | ···················· | ···················· | ···················· | ···················· | ···················· | ···················· | |
| H | 3449 hCoV-19/Indonesia/JK-NIHRD-MI41613/2020\|EPI_ISL_53 | ···················· | ···················· | ···················· | ···················· | ···················· | ···················· | |
| H | 3450 hCoV-19/Indonesia/JK-NIHRD-MI52885/2020\|EPI_ISL_53 | ···················· | ···················· | ···················· | ···················· | ···················· | ···················· | |
| H | 3451 hCoV-19/Indonesia/PA-NIHRD-C0710717/2020\|EPI_ISL_5 | ···················· | ···················· | ···················· | ···················· | ···················· | ···················· | |
| H | 3452 hCoV-19/Indonesia/PA-NIHRD-C0710684/2020\|EPI_ISL_5 | ···················· | ···················· | ···················· | ···················· | ···················· | ···················· | |
| H | 3453 hCoV-19/Indonesia/JK-NIHRD-MI2851/2020\|EPI_ISL_538 | ···················· | ···················· | ···················· | ···················· | ···················· | ···················· | |
| H | 3454 hCoV-19/Indonesia/JK-NIHRD-MI52946/2020\|EPI_ISL_53 | ···················· | ···················· | ···················· | ···················· | ···················· | ···················· | |
| H | 3455 hCoV-19/Indonesia/KS-NIHRD-PME4900/2020\|EPI_ISL_53 | ···················· | ···················· | ···················· | ···················· | ···················· | ···················· | |
| H | 3456 hCoV-19/Indonesia/JI-NIHRD-PME5011/2020\|EPI_ISL_53 | ···················· | ·A·················· | ···················· | ···················· | ···················· | ···················· | |
| H | 3457 hCoV-19/Indonesia/PA-NIHRD-PME4306/2020\|EPI_ISL_53 | ···················· | ···················· | ···················· | ···················· | ···················· | ···················· | |
| H | 3458 hCoV-19/Indonesia/JB-NIHRD-PME4992/2020\|EPI_ISL_53 | ···················· | ···A················ | ···················· | ···················· | ···················· | ···················· | |
| H | 3459 hCoV-19/USA/NC-SLPH-0001/2020\|EPI_ISL_538514\|2020- | ···················· | ···················· | ···················· | ···················· | ···················· | ···················· | |
| H | 3460 hCoV-19/USA/NC-SLPH-0002/2020\|EPI_ISL_538515\|2020- | ···················· | ···················· | ···················· | ···················· | ···················· | ···················· | |
| H | 3461 hCoV-19/USA/NC-SLPH-0003/2020\|EPI_ISL_538516\|2020- | ···················· | ···················· | ···················· | ···················· | ···················· | ···················· | |
| H | 3462 hCoV-19/USA/NC-SLPH-0007/2020\|EPI_ISL_538517\|2020- | ···················· | ···················· | ···········T········ | ···················· | ···················· | ···················· | |
| H | 3463 hCoV-19/USA/NC-SLPH-0009/2020\|EPI_ISL_538519\|2020- | ···················· | ···················· | ···········T········ | ···················· | ···················· | ···················· | |
| H | 3464 hCoV-19/USA/NC-SLPH-0011/2020\|EPI_ISL_538520\|2020- | ···················· | ·A·················· | ···················· | ···················· | ···················· | ···················· | |
| H | 3465 hCoV-19/USA/NC-SLPH-0017/2020\|EPI_ISL_538521\|2020- | ···················· | ···················· | ···················· | ···················· | ···················· | ···················· | |
| H | 3466 hCoV-19/USA/NC-SLPH-0018/2020\|EPI_ISL_538522\|2020- | ···················· | ···················· | ···················· | ···················· | ···················· | ···················· | |
| H | 3467 hCoV-19/USA/NC-SLPH-0010/2020\|EPI_ISL_538523\|2020- | ···················· | ···················· | ···················· | ···················· | ···················· | ···················· | |
| H | 3468 hCoV-19/USA/WY-WYPHL-20010570/2020\|EPI_ISL_539800\| | ···················· | ···················· | ···················· | ···················· | ···················· | ···················· | |
| H | 3469 hCoV-19/USA/WY-WYPHL-20010697/2020\|EPI_ISL_539801\| | ···················· | ···················· | ···················· | ···················· | ···················· | ···················· | |
| H | 3470 hCoV-19/USA/WY-WYPHL-20011372/2020\|EPI_ISL_539802\| | ···················· | ···················· | ···················· | ···················· | ···················· | ···················· | |
| H | 3471 hCoV-19/Hong Kong/VM20061555/2020\|EPI_ISL_539803\|2 | ···················· | ···················· | ···················· | ···················· | ···················· | ···················· | |
| H | 3472 hCoV-19/Hong Kong/VM20030365-2/2020\|EPI_ISL_539804 | ···················· | ···················· | ···················· | ···················· | ···················· | ···················· | |
| H | 3473 hCoV-19/Hong Kong/VM20030491-2/2020\|EPI_ISL_539805 | ···················· | ···················· | ···················· | ···················· | ···················· | ···················· | |
| H | 3474 hCoV-19/Hong Kong/VM20031161/2020\|EPI_ISL_539806\|2 | ···················· | ···················· | ···················· | ···················· | ···················· | ···················· | |
| H | 3475 hCoV-19/Hong Kong/VM20030723/2020\|EPI_ISL_539807\|2 | ···················· | ···················· | ···················· | ···················· | ···················· | ···················· | |
| H | 3476 hCoV-19/Hong Kong/VM20031162/2020\|EPI_ISL_539808\|2 | ···················· | ···················· | ···················· | ···················· | ···················· | ···················· | |
| H | 3477 hCoV-19/Hong Kong/VM20040795/2020\|EPI_ISL_539809\|2 | ···················· | ···················· | ···················· | ···················· | ···················· | ···················· | |
| H | 3478 hCoV-19/Hong Kong/VM20059282/2020\|EPI_ISL_539810\|2 | ···················· | ···················· | ···················· | ···················· | ···················· | ···················· | |
| H | 3479 hCoV-19/Hong Kong/VM20060398/2020\|EPI_ISL_539811\|2 | ···················· | ·A·················· | ···················· | ···················· | ···················· | ··A················· | |
| H | 3480 hCoV-19/Hong Kong/VM20060346/2020\|EPI_ISL_539812\|2 | ···················· | ···················· | ···················· | ···················· | ···················· | ···················· | |
| H | 3481 hCoV-19/Hong Kong/VM20063264/2020\|EPI_ISL_539813\|2 | ···················· | ···················· | ···················· | ···················· | ···················· | ···················· | |
| H | 3482 hCoV-19/Hong Kong/VM20066524/2020\|EPI_ISL_539814\|2 | ···················· | ···················· | ···················· | ···················· | ···················· | ···················· | |
| H | 3483 hCoV-19/Hong Kong/VM20066568/2020\|EPI_ISL_539815\|2 | ···················· | ···················· | ···················· | ···················· | ···················· | ···················· | |
| H | 3484 hCoV-19/Hong Kong/VM20106598/2020\|EPI_ISL_539816\|2 | ···················· | ···················· | ···················· | ···················· | ···················· | ···················· | |
| H | 3485 hCoV-19/Hong Kong/VM20108307/2020\|EPI_ISL_539817\|2 | ···················· | ···················· | ···················· | ···················· | ···················· | ···················· | |
| H | 3486 hCoV-19/Hong Kong/VM20107470/2020\|EPI_ISL_539818\|2 | ···················· | ···················· | ···················· | ···················· | ···················· | ···················· | |
| H | 3487 hCoV-19/Hong Kong/VM20107468/2020\|EPI_ISL_539819\|2 | ···················· | ···················· | ···················· | ···················· | ···················· | ···················· | |
| H | 3488 hCoV-19/Hong Kong/VM20006777/2020\|EPI_ISL_539820\|2 | ···················· | ···················· | ···················· | ···················· | ···················· | ···················· | |
| H | 3489 hCoV-19/Hong Kong/VM20108924/2020\|EPI_ISL_539821\|2 | ···················· | ···················· | ···················· | ···················· | ···················· | ···················· | |
| H | 3490 hCoV-19/Hong Kong/VB20175873/2020\|EPI_ISL_539822\|2 | ···················· | ···················· | ···················· | ···················· | ···················· | ···················· | |
| H | 3491 hCoV-19/Hong Kong/VB20175856/2020\|EPI_ISL_539823\|2 | ···················· | ···················· | ···················· | ···················· | ···················· | ···················· | |

| | | | | | | | | |
|---|---|---|---|---|---|---|---|---|
| H | 3492 hCoV-19/USA/MN-MDH-1630/2020\|EPI_ISL_539824\|2020-0 | ···················· | ···················· | ···················· | ···················· | ···················· | ···················· | |
| H | 3493 hCoV-19/USA/MN-MDH-1631/2020\|EPI_ISL_539825\|2020-0 | ···················· | ···················· | ···················· | ···················· | ···················· | ···················· | |
| H | 3494 hCoV-19/USA/MN-MDH-1632/2020\|EPI_ISL_539826\|2020-0 | ···················· | ···················· | ···················· | ···················· | ···················· | ···················· | |
| H | 3495 hCoV-19/USA/MN-MDH-1633/2020\|EPI_ISL_539827\|2020-0 | ···················· | ···················· | ···················· | ···················· | ···················· | ···················· | |
| H | 3496 hCoV-19/USA/MN-MDH-1634/2020\|EPI_ISL_539828\|2020-0 | ···················· | ···················· | ···················· | ···················· | ···················· | ···················· | |
| H | 3497 hCoV-19/USA/MN-MDH-1635/2020\|EPI_ISL_539829\|2020-0 | ···················· | ···················· | ···················· | ···················· | ···················· | ···················· | |
| H | 3498 hCoV-19/USA/MN-MDH-1636/2020\|EPI_ISL_539830\|2020-0 | ···················· | ···················· | ···················· | ···················· | ···················· | ···················· | |
| H | 3499 hCoV-19/USA/MN-MDH-1637/2020\|EPI_ISL_539831\|2020-0 | ···················· | ···················· | ···················· | ···················· | ···················· | ···················· | |
| H | 3500 hCoV-19/USA/MN-MDH-1638/2020\|EPI_ISL_539832\|2020-0 | ···················· | ···················· | ···················· | ···················· | ···················· | ···················· | |
| H | 3501 hCoV-19/USA/MN-MDH-1639/2020\|EPI_ISL_539833\|2020-0 | ···················· | ···················· | ···················· | ···················· | ···················· | ···················· | |
| H | 3502 hCoV-19/USA/MN-MDH-1640/2020\|EPI_ISL_539834\|2020-0 | ···················· | ···················· | ···················· | ···················· | ···················· | ···················· | |
| H | 3503 hCoV-19/USA/MN-MDH-1641/2020\|EPI_ISL_539835\|2020-0 | ···················· | ···················· | ···················· | ···················· | ···················· | ···················· | |
| H | 3504 hCoV-19/USA/MN-MDH-1642/2020\|EPI_ISL_539836\|2020-0 | ···················· | ···················· | ···················· | ···················· | ···················· | ···················· | |
| H | 3505 hCoV-19/USA/MN-MDH-1643/2020\|EPI_ISL_539837\|2020-0 | ···················· | ···················· | ···················· | ···················· | ···················· | ···················· | |
| H | 3506 hCoV-19/USA/MN-MDH-1644/2020\|EPI_ISL_539838\|2020-0 | ···················· | ···················· | ···················· | ···················· | ···················· | ···················· | |
| H | 3507 hCoV-19/USA/MN-MDH-1645/2020\|EPI_ISL_539839\|2020-0 | ···················· | ···················· | ···················· | ···················· | ···················· | ···················· | |
| H | 3508 hCoV-19/USA/MN-MDH-1646/2020\|EPI_ISL_539840\|2020-0 | ···················· | ···················· | ···················· | ···················· | ···················· | ···················· | |
| H | 3509 hCoV-19/USA/MN-MDH-1647/2020\|EPI_ISL_539841\|2020-0 | ···················· | ···················· | ···················· | ···················· | ···················· | ···················· | |
| H | 3510 hCoV-19/USA/MN-MDH-1648/2020\|EPI_ISL_539842\|2020-0 | ···················· | ·············A······ | ···················· | ···················· | ···················· | ···················· | |
| H | 3511 hCoV-19/USA/MN-MDH-1649/2020\|EPI_ISL_539843\|2020-0 | ···················· | ···················· | ···················· | ···················· | ···················· | ···················· | |
| H | 3512 hCoV-19/USA/MN-MDH-1650/2020\|EPI_ISL_539844\|2020-0 | ···················· | ···················· | ···················· | ···················· | ···················· | ···················· | |
| H | 3513 hCoV-19/USA/MN-MDH-1651/2020\|EPI_ISL_539845\|2020-0 | ········T··········· | ···················· | ···················· | ···················· | ···················· | ···················· | |
| H | 3514 hCoV-19/USA/MN-MDH-1652/2020\|EPI_ISL_539846\|2020-0 | ···················· | ···················· | ···················· | ···················· | ···················· | ···················· | |
| H | 3515 hCoV-19/USA/MN-MDH-1653/2020\|EPI_ISL_539847\|2020-0 | ···················· | ·A·················· | ···················· | ···················· | ···················· | ···················· | |
| H | 3516 hCoV-19/USA/MN-MDH-1654/2020\|EPI_ISL_539848\|2020-0 | ···················· | ···················· | ···················· | ···················· | ···················· | ···················· | |
| H | 3517 hCoV-19/USA/MN-MDH-1655/2020\|EPI_ISL_539849\|2020-0 | ···················· | ···················· | ···················· | ···················· | ···················· | ···················· | |
| H | 3518 hCoV-19/Hong Kong/VM20005737-2/2020\|EPI_ISL_539850 | ···················· | ···················· | ···················· | ···················· | ···················· | ···················· | |
| H | 3519 hCoV-19/Hong Kong/VM20008964-2/2020\|EPI_ISL_539851 | ···················· | ···················· | ···················· | ···················· | ···················· | ···················· | |
| H | 3520 hCoV-19/Scotland/CVR4108/2020\|EPI_ISL_540810\|2020- | ···················· | ···················· | ···················· | ···················· | ···················· | ···················· | |
| H | 3521 hCoV-19/Scotland/CVR4133/2020\|EPI_ISL_540824\|2020- | ···················· | ···················· | ···················· | ···················· | ···················· | ···················· | |
| H | 3522 hCoV-19/Scotland/CVR4135/2020\|EPI_ISL_540825\|2020- | ···················· | ···················· | ···················· | ···················· | ···················· | ···················· | |
| H | 3523 hCoV-19/Scotland/CVR4136/2020\|EPI_ISL_540826\|2020- | ···················· | ···················· | ···················· | ···················· | ···················· | ···················· | |
| H | 3524 hCoV-19/Scotland/CVR4139/2020\|EPI_ISL_540827\|2020- | ···················· | ···················· | ···················· | ···················· | ···················· | ···················· | |
| H | 3525 hCoV-19/Scotland/CVR4140/2020\|EPI_ISL_540828\|2020- | ···················· | ·A·················· | ···················· | ···················· | ···················· | ··A················· | |
| H | 3526 hCoV-19/Scotland/CVR4141/2020\|EPI_ISL_540829\|2020- | ···················· | ···················· | ···················· | ···················· | ···················· | ···················· | |
| H | 3527 hCoV-19/Scotland/CVR4142/2020\|EPI_ISL_540830\|2020- | ···················· | ···················· | ···················· | ···················· | ···················· | ···················· | |
| H | 3528 hCoV-19/Scotland/CVR4143/2020\|EPI_ISL_540831\|2020- | ···················· | ···················· | ···················· | ···················· | ···················· | ···················· | |
| H | 3529 hCoV-19/Scotland/CVR4144/2020\|EPI_ISL_540832\|2020- | ···················· | ···················· | ···················· | ···················· | ···················· | ···················· | |
| H | 3530 hCoV-19/Scotland/CVR4146/2020\|EPI_ISL_540833\|2020- | ···················· | ···················· | ···················· | ···················· | ···················· | ···················· | |
| H | 3531 hCoV-19/Scotland/CVR4149/2020\|EPI_ISL_540834\|2020- | ···················· | ···················· | ···················· | ···················· | ···················· | ···················· | |
| H | 3532 hCoV-19/Scotland/CVR4150/2020\|EPI_ISL_540835\|2020- | ···················· | ···················· | ···················· | ···················· | ···················· | ···················· | |
| H | 3533 hCoV-19/Scotland/CVR4151/2020\|EPI_ISL_540836\|2020- | ···················· | ···················· | ···················· | ···················· | ···················· | ···················· | |
| H | 3534 hCoV-19/Scotland/CVR4154/2020\|EPI_ISL_540839\|2020- | ···················· | ···················· | ···················· | ···················· | ···················· | ···················· | |
| H | 3536 hCoV-19/USA/TX-HMH-3582/2020\|EPI_ISL_543040\|2020-0 | ···················· | ···················· | ···················· | ···················· | ···················· | ···················· | |
| H | 3544 hCoV-19/USA/TX-HMH-3594/2020\|EPI_ISL_543050\|2020-0 | ···················· | ·············A······ | ···················· | ···················· | ···················· | ···················· | |
| H | 3548 hCoV-19/USA/TX-HMH-3599/2020\|EPI_ISL_543055\|2020-0 | ···················· | ···················· | ···················· | ···················· | ···················· | ···················· | |
| H | 3553 hCoV-19/USA/TX-HMH-3607/2020\|EPI_ISL_543062\|2020-0 | ···················· | ···················· | ···················· | ···················· | ···················· | ··A················· | |
| H | 3554 hCoV-19/USA/TX-HMH-3608/2020\|EPI_ISL_543063\|2020-0 | ···················· | ···················· | ···················· | ···················· | ···················· | ···················· | |
| H | 3555 hCoV-19/USA/TX-HMH-3609/2020\|EPI_ISL_543064\|2020-0 | ···················· | ···················· | ···················· | ···················· | ···················· | ···················· | |
| H | 3556 hCoV-19/USA/TX-HMH-3610/2020\|EPI_ISL_543065\|2020-0 | ···················· | ···················· | ···················· | ···················· | ···················· | ···················· | |
| H | 3560 hCoV-19/USA/TX-HMH-3617/2020\|EPI_ISL_543070\|2020-0 | ···················· | ···················· | ···················· | ···················· | N··················· | NN·N················ | 1 |
| H | 3561 hCoV-19/USA/TX-HMH-3618/2020\|EPI_ISL_543071\|2020-0 | ···················· | ···················· | ···················· | ···················· | ···················· | ···················· | |
| H | 3562 hCoV-19/USA/TX-HMH-3619/2020\|EPI_ISL_543072\|2020-0 | ···················· | ···················· | ···················· | ···················· | ···················· | ···················· | |
| H | 3563 hCoV-19/USA/TX-HMH-3620/2020\|EPI_ISL_543073\|2020-0 | ···················· | ···················· | ···················· | ···················· | ···················· | ···················· | |
| H | 3565 hCoV-19/USA/TX-HMH-3623/2020\|EPI_ISL_543076\|2020-0 | ···················· | ···················· | ···················· | ···················· | ···················· | ···················· | |
| H | 3566 hCoV-19/USA/TX-HMH-3624/2020\|EPI_ISL_543077\|2020-0 | ···················· | ···················· | ···················· | ···················· | ···················· | ···················· | |
| H | 3567 hCoV-19/USA/TX-HMH-3626/2020\|EPI_ISL_543078\|2020-0 | ···················· | ···················· | ···················· | ···················· | ···················· | ···················· | |
| H | 3568 hCoV-19/USA/TX-HMH-3627/2020\|EPI_ISL_543079\|2020-0 | ···················· | ···················· | ···················· | ···················· | ···················· | ···················· | |
| H | 3572 hCoV-19/USA/TX-HMH-3635/2020\|EPI_ISL_543085\|2020-0 | ···················· | ·A·················· | ···················· | ···················· | ···················· | ···················· | |
| H | 3574 hCoV-19/USA/TX-HMH-3639/2020\|EPI_ISL_543087\|2020-0 | ···················· | ···················· | ···················· | ···················· | ···················· | ···················· | |
| H | 3575 hCoV-19/USA/TX-HMH-3640/2020\|EPI_ISL_543088\|2020-0 | ···················· | ···················· | ···················· | ···················· | ···················· | ···················· | |
| H | 3577 hCoV-19/USA/TX-HMH-3644/2020\|EPI_ISL_543090\|2020-0 | ···················· | ·A·················· | ···················· | ···················· | ···················· | ···················· | |
| H | 3578 hCoV-19/USA/TX-HMH-3645/2020\|EPI_ISL_543091\|2020-0 | ···················· | ···················· | ···················· | ···················· | ···················· | ···················· | |
| H | 3579 hCoV-19/USA/TX-HMH-3646/2020\|EPI_ISL_543092\|2020-0 | ···················· | ···················· | ···················· | ···················· | ···················· | ···················· | |
| H | 3580 hCoV-19/USA/TX-HMH-3647/2020\|EPI_ISL_543093\|2020-0 | ···················· | ···················· | ···················· | ···················· | ···················· | ···················· | |
| H | 3584 hCoV-19/USA/TX-HMH-3654/2020\|EPI_ISL_543099\|2020-0 | ···················· | ···················· | ···················· | ···················· | ···················· | ···················· | |
| H | 3585 hCoV-19/England/QEUH-9697AD/2020\|EPI_ISL_531000\|20 | ···················· | ···················· | ···················· | ···················· | ···················· | ···················· | |
| H | 3586 hCoV-19/England/QEUH-9655D4/2020\|EPI_ISL_531001\|20 | ···················· | ···················· | ···················· | ···················· | ···················· | ···················· | |
| H | 3587 hCoV-19/Scotland/QEUH-964E16/2020\|EPI_ISL_531002\|2 | ···················· | ···················· | ···················· | ···················· | ···················· | ···················· | |
| H | 3588 hCoV-19/England/QEUH-965264/2020\|EPI_ISL_531003\|20 | ···················· | ···················· | ···················· | ···················· | ···················· | ···················· | |
| H | 3589 hCoV-19/Scotland/QEUH-964A5E/2020\|EPI_ISL_531004\|2 | ···················· | ···················· | ···················· | ···················· | ···················· | ···················· | |
| H | 3590 hCoV-19/England/QEUH-965668/2020\|EPI_ISL_531005\|20 | ···················· | ·A·················· | ···················· | ···················· | ···················· | ··A················· | |
| H | 3591 hCoV-19/Scotland/QEUH-96549B/2020\|EPI_ISL_531006\|2 | ···················· | ···················· | ···················· | ···················· | ···················· | ···················· | |
| H | 3592 hCoV-19/England/QEUH-965219/2020\|EPI_ISL_531007\|20 | ···················· | ···················· | ···················· | ···················· | ···················· | ···················· | |
| H | 3593 hCoV-19/England/QEUH-9653F8/2020\|EPI_ISL_531009\|20 | ···················· | ···················· | ···················· | ···················· | ···················· | ···················· | |
| H | 3594 hCoV-19/England/QEUH-9658CC/2020\|EPI_ISL_531010\|20 | ···················· | ···················· | ···················· | ···················· | ···················· | ···················· | |
| H | 3595 hCoV-19/England/QEUH-965835/2020\|EPI_ISL_531011\|20 | ···················· | ···················· | ···················· | ···················· | ···················· | ···················· | |
| H | 3596 hCoV-19/England/QEUH-9651EF/2020\|EPI_ISL_531013\|20 | ···················· | ···················· | ···················· | ···················· | ···················· | ···················· | |
| H | 3597 hCoV-19/England/QEUH-96978F/2020\|EPI_ISL_531014\|20 | ···················· | ···················· | ···················· | ···················· | ···················· | ···················· | |
| H | 3598 hCoV-19/England/QEUH-9652DD/2020\|EPI_ISL_531015\|20 | ···················· | ···················· | ···················· | ···················· | ···················· | ···················· | |

```
H    3599 hCoV-19/England/QEUH-964DFB/2020|EPI_ISL_531016|20   ···················· ···················· ···················· ···················· ···················· ····················
H    3600 hCoV-19/Scotland/QEUH-964CB2/2020|EPI_ISL_531017|2   ···················· ···················· ···················· ···················· ···················· ····················
H    3601 hCoV-19/England/QEUH-96520A/2020|EPI_ISL_531018|20   ···················· ···················· ···················· ···················· ···················· ····················
H    3602 hCoV-19/Scotland/QEUH-965765/2020|EPI_ISL_531020|2   ···················· ···················· ···················· ···················· ···················· ····················
H    3603 hCoV-19/England/QEUH-969716/2020|EPI_ISL_531021|20   ···················· ···················· ···················· ···················· ···················· ····················
H    3604 hCoV-19/England/QEUH-964D19/2020|EPI_ISL_531022|20   ···················· ···················· ···················· ···················· ···················· ····················
H    3605 hCoV-19/England/QEUH-969B65/2020|EPI_ISL_531023|20   ···················· ···················· ···················· ···················· ···················· ····················
H    3606 hCoV-19/England/QEUH-9656FF/2020|EPI_ISL_531024|20   ···················· ···················· ···················· ···················· ···················· ····················
H    3607 hCoV-19/England/QEUH-965950/2020|EPI_ISL_531026|20   ···················· ···················· ···················· ···················· ···················· ····················
H    3608 hCoV-19/England/QEUH-969AE0/2020|EPI_ISL_531027|20   ···················· ···················· ···················· ···················· ···················· ····················
H    3609 hCoV-19/England/QEUH-9698E6/2020|EPI_ISL_531028|20   ···················· ···················· ···················· ···················· ···················· ····················
H    3610 hCoV-19/Northern Ireland/QEUH-969761/2020|EPI_ISL_   ···················· ···················· ···················· ···················· ···················· ····················
H    3611 hCoV-19/Scotland/QEUH-9697E9/2020|EPI_ISL_531032|2   ···················· ···················· ···················· ···················· ···················· ····················
H    3612 hCoV-19/Scotland/QEUH-96986E/2020|EPI_ISL_531033|2   ···················· ···················· ···················· ···················· ···················· ····················
H    3613 hCoV-19/England/QEUH-96503D/2020|EPI_ISL_531034|20   ···················· ···················· ···················· ···················· ···················· ····················
H    3614 hCoV-19/England/QEUH-964C76/2020|EPI_ISL_531035|20   ···················· ···················· ···················· ···················· ······T············· ····················
H    3615 hCoV-19/England/QEUH-964C1C/2020|EPI_ISL_531036|20   ···················· ···················· ···················· ···················· ···················· ····················
H    3616 hCoV-19/England/QEUH-964CA3/2020|EPI_ISL_531037|20   ···················· ···················· ···················· ···················· ···················· ····················
H    3617 hCoV-19/Scotland/QEUH-964AB8/2020|EPI_ISL_531038|2   ···················· ···················· ···················· ···················· ···················· ····················
H    3618 hCoV-19/England/QEUH-9653AD/2020|EPI_ISL_531039|20   ···················· ···················· ···················· ···················· ···················· ····················
H    3619 hCoV-19/Scotland/QEUH-969707/2020|EPI_ISL_531040|2   ···················· ···················· ···················· ···················· ···················· ····················
H    3620 hCoV-19/England/QEUH-9649AC/2020|EPI_ISL_531041|20   ···················· ·A·················· ···················· ···················· ···················· ···A················
H    3621 hCoV-19/England/QEUH-964BE2/2020|EPI_ISL_531042|20   ···················· ···················· ···················· ···················· ···················· ····················
H    3622 hCoV-19/Northern Ireland/QEUH-96505B/2020|EPI_ISL_   ···················· ···················· ···················· ···················· ···················· ····················
H    3623 hCoV-19/England/QEUH-9656E0/2020|EPI_ISL_531044|20   ···················· ···················· ···················· ···················· ···················· ····················
H    3624 hCoV-19/Scotland/QEUH-965871/2020|EPI_ISL_531045|2   ···················· ···················· ···················· ···················· ···················· ····················
H    3625 hCoV-19/England/QEUH-964DDD/2020|EPI_ISL_531046|20   ···················· ···················· ···················· ···················· ···················· ····················
H    3626 hCoV-19/Northern Ireland/QEUH-964F6E/2020|EPI_ISL_   ···················· ···················· ···················· ···················· ···················· ····················
H    3627 hCoV-19/Scotland/QEUH-9697DA/2020|EPI_ISL_531048|2   ···················· ···················· ···················· ···················· ···················· ····················
H    3628 hCoV-19/Scotland/QEUH-965079/2020|EPI_ISL_531049|2   ···················· ···················· ···················· ···················· ···················· ····················
H    3629 hCoV-19/England/QEUH-965325/2020|EPI_ISL_531050|20   ···················· ···················· ···················· ···················· ···················· ····················
H    3630 hCoV-19/Scotland/QEUH-964AD6/2020|EPI_ISL_531052|2   ···················· ···················· ···················· ···················· ···················· ····················
H    3631 hCoV-19/Northern Ireland/QEUH-964F40/2020|EPI_ISL_   ···················· ···················· ···················· ···················· ···················· ····················
H    3632 hCoV-19/Scotland/QEUH-969840/2020|EPI_ISL_531054|2   ···················· ···················· ···················· ···················· ···················· ····················
H    3633 hCoV-19/Northern Ireland/QEUH-9699F2/2020|EPI_ISL_   ···················· ···················· ···················· ···················· ···················· ····················
H    3634 hCoV-19/Scotland/QEUH-9696CE/2020|EPI_ISL_531057|2   ···················· ···················· ···················· ···················· ···················· ····················
H    3635 hCoV-19/England/QEUH-969B83/2020|EPI_ISL_531058|20   ···················· ···················· ···················· ···················· ···················· ····················
H    3636 hCoV-19/England/QEUH-964C2B/2020|EPI_ISL_531059|20   ···················· ···················· ···················· ···················· ···················· ····················
H    3637 hCoV-19/Scotland/QEUH-95EA48/2020|EPI_ISL_532321|2   ···················· ···················· ···················· ···················· ···················· ····················
H    3638 hCoV-19/England/QEUH-95ED4F/2020|EPI_ISL_532322|20   ···················· ···················· ···················· ···················· ···················· ····················
H    3639 hCoV-19/England/QEUH-95F86B/2020|EPI_ISL_532323|20   ···················· ···················· ···················· ···················· ···················· ····················
H    3640 hCoV-19/England/QEUH-95E5DE/2020|EPI_ISL_532324|20   ···················· ···················· ···················· ···················· ···················· ····················
H    3641 hCoV-19/England/QEUH-95F7F5/2020|EPI_ISL_532325|20   ···················· ···················· ···················· ···················· ···················· ····················
H    3642 hCoV-19/Northern Ireland/QEUH-95EAA2/2020|EPI_ISL_   ···················· ···················· ···················· ···················· ···················· ····················
H    3643 hCoV-19/Scotland/QEUH-95F027/2020|EPI_ISL_532328|2   ···················· ·A·················· ···················· ···················· ···················· ····················
H    3644 hCoV-19/England/QEUH-95F634/2020|EPI_ISL_532329|20   ···················· ···················· ···················· ···················· ···················· ····················
H    3645 hCoV-19/England/QEUH-95E94B/2020|EPI_ISL_532330|20   ···················· ···················· ···················· ···················· ···················· ····················
H    3646 hCoV-19/England/QEUH-95EE6A/2020|EPI_ISL_532331|20   ···················· ···················· ···················· ···················· ···················· ····················
H    3647 hCoV-19/England/QEUH-95F625/2020|EPI_ISL_532333|20   ···················· ···················· ···················· ···················· ···················· ····················
H    3648 hCoV-19/Scotland/QEUH-95F27C/2020|EPI_ISL_532334|2   ···················· ···················· ···················· ···················· ···················· ····················
H    3649 hCoV-19/Scotland/QEUH-95F9A4/2020|EPI_ISL_532335|2   ···················· ···················· ···················· ···················· ···················· ····················
H    3650 hCoV-19/England/QEUH-95F8B6/2020|EPI_ISL_532336|20   ···················· ···················· ···················· ···················· ···················· ····················
H    3651 hCoV-19/Northern Ireland/QEUH-95EB27/2020|EPI_ISL_   ···················· ···················· ···················· ···················· ···················· ····················
H    3652 hCoV-19/Scotland/QEUH-95E705/2020|EPI_ISL_532338|2   ···················· ···················· ···················· ···················· ···················· ····················
H    3653 hCoV-19/England/QEUH-95FC23/2020|EPI_ISL_532339|20   ···················· ···················· ···················· ···················· ···················· ····················
H    3654 hCoV-19/Scotland/QEUH-95F9E0/2020|EPI_ISL_532340|2   ···················· ···················· ···················· ···················· ···················· ····················
H    3655 hCoV-19/England/QEUH-95E811/2020|EPI_ISL_532341|20   K··················· ···················· ···················· ···················· ···················· ····················
H    3656 hCoV-19/England/QEUH-95F5FB/2020|EPI_ISL_532343|20   ···················· ···················· ···················· ···················· ···················· ····················
H    3657 hCoV-19/England/QEUH-95F0CD/2020|EPI_ISL_532344|20   ···················· ···················· ···················· ···················· ···················· ····················
H    3658 hCoV-19/Scotland/QEUH-95EF3A/2020|EPI_ISL_532347|2   ···················· ···················· ···················· ···················· ···················· ····················
H    3659 hCoV-19/England/QEUH-95E69F/2020|EPI_ISL_532348|20   ···················· ···················· ···················· ···················· ···················· ····················
H    3660 hCoV-19/Scotland/QEUH-95E90F/2020|EPI_ISL_532349|2   ···················· ···················· ···················· ···················· ···················· ····················
H    3661 hCoV-19/Northern Ireland/QEUH-95EEE2/2020|EPI_ISL_   ···················· ···················· ···················· ···················· ···················· ····················
H    3662 hCoV-19/Scotland/QEUH-95FA92/2020|EPI_ISL_532351|2   ···················· ···················· ···················· ···················· ···················· ····················
H    3663 hCoV-19/England/QEUH-95FA83/2020|EPI_ISL_532352|20   ···················· ···················· ···················· ···················· ···················· ····················
H    3664 hCoV-19/Scotland/QEUH-95F9FF/2020|EPI_ISL_532353|2   ···················· ···················· ···················· ···················· ···················· ····················
H    3665 hCoV-19/England/QEUH-95E8D5/2020|EPI_ISL_532354|20   ···················· ···················· ···················· ···················· ···················· ····················
H    3666 hCoV-19/Scotland/QEUH-95FB71/2020|EPI_ISL_532355|2   ···················· ···················· ···················· ···················· ···················· ····················
H    3667 hCoV-19/Scotland/QEUH-95EE97/2020|EPI_ISL_532356|2   ···················· ···················· ···················· ···················· ···················· ····················
H    3668 hCoV-19/England/QEUH-95EFD0/2020|EPI_ISL_532357|20   ···················· ···················· ···················· ···················· ···················· ····················
H    3669 hCoV-19/England/QEUH-95EF76/2020|EPI_ISL_532359|20   ···················· ···················· ···················· ···················· ···················· ····················
H    3670 hCoV-19/Northern Ireland/QEUH-95EB36/2020|EPI_ISL_   ···················· ···················· ···················· ···················· ···················· ····················
H    3671 hCoV-19/Northern Ireland/QEUH-95FAA1/2020|EPI_ISL_   ···················· ···················· ···················· ···················· ···················· ····················
H    3672 hCoV-19/Scotland/QEUH-95E617/2020|EPI_ISL_532363|2   ···················· ···················· ···················· ···················· ···················· ····················
H    3673 hCoV-19/Scotland/QEUH-95E8C6/2020|EPI_ISL_532364|2   ···················· ···················· ···················· ···················· ···················· ····················
H    3674 hCoV-19/England/QEUH-95FB08/2020|EPI_ISL_532366|20   ···················· ···················· ···················· ···················· ···················· ····················
H    3675 hCoV-19/Northern Ireland/QEUH-95EA66/2020|EPI_ISL_   ···················· ···················· ···················· ···················· ···················· ····················
H    3676 hCoV-19/Northern Ireland/QEUH-95FA56/2020|EPI_ISL_   ···················· ···················· ···················· ···················· ···················· ····················
H    3677 hCoV-19/England/QEUH-95FBF9/2020|EPI_ISL_532369|20   ···················· ···················· ···················· ···················· ···················· ····················
H    3678 hCoV-19/Northern Ireland/QEUH-95EF0D/2020|EPI_ISL_   ···················· ···················· ···················· ···················· ···················· ····················
```

| | | | | | | | | |
|---|---|---|---|---|---|---|---|---|
| H | 3679 | hCoV-19/England/QEUH-95E644/2020\|EPI_ISL_532372\|20 | ···················· | ···················· | ···················· | ···················· | ···················· | ···················· |
| H | 3680 | hCoV-19/England/QEUH-95ED9A/2020\|EPI_ISL_532373\|20 | ···················· | ···················· | ···················· | ···················· | ···················· | ···················· |
| H | 3681 | hCoV-19/England/QEUH-95F670/2020\|EPI_ISL_532374\|20 | ···················· | ···················· | ···················· | ···················· | ···················· | ···················· |
| H | 3682 | hCoV-19/England/QEUH-95F4C1/2020\|EPI_ISL_532375\|20 | ···················· | ···················· | ···················· | ···················· | ···················· | ···················· |
| H | 3683 | hCoV-19/England/QEUH-95E76F/2020\|EPI_ISL_532377\|20 | ···················· | ···················· | ···················· | ···················· | ···················· | ···················· |
| H | 3684 | hCoV-19/Wales/QEUH-95F69E/2020\|EPI_ISL_532378\|2020 | ···················· | ···················· | ···················· | ···················· | ···················· | ···················· |
| H | 3685 | hCoV-19/England/QEUH-95E7E7/2020\|EPI_ISL_532380\|20 | ···················· | ···················· | ···················· | ···················· | ···················· | ···················· |
| H | 3686 | hCoV-19/Northern Ireland/QEUH-95E9A5/2020\|EPI_ISL_ | ···················· | ···················· | ···················· | ···················· | ···················· | ···················· |
| H | 3687 | hCoV-19/England/QEUH-95F91D/2020\|EPI_ISL_532382\|20 | ···················· | ···················· | ···················· | ···················· | ···················· | ···················· |
| H | 3688 | hCoV-19/England/QEUH-95F467/2020\|EPI_ISL_532383\|20 | ···················· | ···················· | ···················· | ···················· | ···················· | ···················· |
| H | 3689 | hCoV-19/Scotland/QEUH-95F9D1/2020\|EPI_ISL_532384\|2 | ···················· | ···················· | ···················· | ···················· | ···················· | ···················· |
| H | 3690 | hCoV-19/England/QEUH-95FA38/2020\|EPI_ISL_532386\|20 | ···················· | ···················· | ···················· | ···················· | ···················· | ···················· |
| H | 3691 | hCoV-19/England/QEUH-95F977/2020\|EPI_ISL_532387\|20 | ···················· | ···················· | ···················· | ···················· | ···················· | ···················· |
| H | 3692 | hCoV-19/Scotland/QEUH-95EA1B/2020\|EPI_ISL_532388\|2 | ···················· | ···················· | ···················· | ···················· | ···················· | ···················· |
| H | 3693 | hCoV-19/England/QEUH-95F063/2020\|EPI_ISL_532389\|20 | ···················· | ···················· | ···················· | ···················· | ···················· | ···················· |
| H | 3694 | hCoV-19/England/QEUH-95F76E/2020\|EPI_ISL_532400\|20 | ···················· | ···················· | ···················· | ···················· | ···················· | ···················· |
| H | 3695 | hCoV-19/Northern Ireland/QEUH-95EAEE/2020\|EPI_ISL_ | ···················· | ···················· | ···················· | ···················· | ···················· | ···················· |
| H | 3696 | hCoV-19/Scotland/QEUH-95EA57/2020\|EPI_ISL_532402\|2 | ···················· | ···················· | ···················· | ···················· | ···················· | ···················· |
| H | 3697 | hCoV-19/England/QEUH-95F968/2020\|EPI_ISL_532403\|20 | ···················· | ···················· | ···················· | ···················· | ···················· | ···················· |
| H | 3698 | hCoV-19/Scotland/QEUH-95F9C2/2020\|EPI_ISL_532404\|2 | ···················· | ···················· | ···················· | ···················· | ···················· | ···················· |
| H | 3699 | hCoV-19/England/QEUH-95E6EA/2020\|EPI_ISL_532406\|20 | ···················· | ···················· | ···················· | ···················· | ···················· | ···················· |
| H | 3700 | hCoV-19/Northern Ireland/QEUH-95EADF/2020\|EPI_ISL_ | ···················· | ···················· | ···················· | ···················· | ···················· | ···················· |
| H | 3701 | hCoV-19/England/QEUH-95E565/2020\|EPI_ISL_532408\|20 | ···················· | ···················· | ···················· | ···················· | ···················· | ···················· |
| H | 3702 | hCoV-19/England/QEUH-95F485/2020\|EPI_ISL_532409\|20 | ···················· | ···················· | ···················· | ···················· | ···················· | ···················· |
| H | 3703 | hCoV-19/England/QEUH-95F2A9/2020\|EPI_ISL_532410\|20 | ···················· | ·A·················· | ···················· | ···················· | ···················· | ···················· |
| H | 3704 | hCoV-19/England/QEUH-95EB18/2020\|EPI_ISL_532411\|20 | ···················· | ···················· | ···················· | ···················· | ···················· | ···················· |
| H | 3705 | hCoV-19/England/QEUH-95F78C/2020\|EPI_ISL_532412\|20 | ···················· | ···················· | ···················· | ···················· | ···················· | ···················· |
| H | 3706 | hCoV-19/England/QEUH-95E556/2020\|EPI_ISL_532413\|20 | ···················· | ···················· | ···················· | ···················· | ···················· | ···················· |
| H | 3707 | hCoV-19/Scotland/QEUH-95ECF7/2020\|EPI_ISL_532414\|2 | ···················· | ···················· | ···················· | ···················· | ···················· | ···················· |
| H | 3708 | hCoV-19/England/QEUH-95FC14/2020\|EPI_ISL_532415\|20 | ···················· | ···················· | ···················· | ···················· | ···················· | ···················· |
| H | 3709 | hCoV-19/Northern Ireland/QEUH-95EDD6/2020\|EPI_ISL_ | ···················· | ···················· | ···················· | ···················· | ···················· | ···················· |
| H | 3710 | hCoV-19/England/QEUH-95EFFE/2020\|EPI_ISL_532417\|20 | ···················· | ···················· | ···················· | ···················· | ···················· | ···················· |
| H | 3711 | hCoV-19/England/QEUH-95EE79/2020\|EPI_ISL_532418\|20 | ···················· | ·A·················· | ···················· | ···················· | ···················· | ··A················· |
| H | 3712 | hCoV-19/Northern Ireland/QEUH-95EBFA/2020\|EPI_ISL_ | ···················· | ···················· | ···················· | ···················· | ···················· | ···················· |
| H | 3713 | hCoV-19/Northern Ireland/QEUH-95EF85/2020\|EPI_ISL_ | ···················· | ···················· | ···················· | ···················· | ···················· | ···················· |
| H | 3714 | hCoV-19/England/QEUH-95F6BC/2020\|EPI_ISL_532421\|20 | ···················· | ···················· | ···················· | ···················· | ···················· | ···················· |
| H | 3715 | hCoV-19/England/OXON-B492E/2020\|EPI_ISL_534983\|202 | ···················· | ···················· | ···················· | ···················· | ···················· | ···················· |
| H | 3716 | hCoV-19/England/OXON-B49B5/2020\|EPI_ISL_534984\|202 | ···················· | ···················· | ···················· | ···················· | ···················· | ···················· |
| H | 3717 | hCoV-19/England/OXON-B49E2/2020\|EPI_ISL_534986\|202 | ···················· | ···················· | ···················· | ···················· | ···················· | ···················· |
| H | 3718 | hCoV-19/England/OXON-B4A2B/2020\|EPI_ISL_534987\|202 | ···················· | ···················· | ···················· | ···················· | ···················· | ···················· |
| H | 3719 | hCoV-19/England/OXON-B4A58/2020\|EPI_ISL_534988\|202 | ···················· | ···················· | ···················· | ···················· | ···················· | ···················· |
| H | 3720 | hCoV-19/England/OXON-B4A67/2020\|EPI_ISL_534989\|202 | ···················· | ···················· | ···················· | ···················· | ···················· | ···················· |
| H | 3721 | hCoV-19/England/OXON-B4A94/2020\|EPI_ISL_534990\|202 | ···················· | ···················· | ···················· | ···················· | ···················· | ···················· |
| H | 3722 | hCoV-19/England/OXON-B4AC1/2020\|EPI_ISL_534993\|202 | ···················· | ···················· | ···················· | ···················· | ···················· | ···················· |
| H | 3723 | hCoV-19/England/OXON-B4B19/2020\|EPI_ISL_534994\|202 | ···················· | ···················· | ···················· | ···················· | ···················· | ···················· |
| H | 3724 | hCoV-19/England/OXON-B4B46/2020\|EPI_ISL_534995\|202 | ···················· | ···················· | ···················· | ···················· | ···················· | ···················· |
| H | 3725 | hCoV-19/England/OXON-B4B73/2020\|EPI_ISL_534996\|202 | ···················· | ···················· | ···················· | ···················· | ···················· | ···················· |
| H | 3726 | hCoV-19/England/OXON-B4B91/2020\|EPI_ISL_534997\|202 | ···················· | ···················· | ···················· | ···················· | ···················· | ···················· |
| H | 3727 | hCoV-19/England/OXON-B4BCE/2020\|EPI_ISL_534998\|202 | ···················· | ···················· | ···················· | ···················· | ···················· | ···················· |
| H | 3728 | hCoV-19/England/QEUH-997E56/2020\|EPI_ISL_537200\|20 | ···················· | ···················· | ···················· | ···················· | ···················· | ···················· |
| H | 3729 | hCoV-19/England/QEUH-997D95/2020\|EPI_ISL_537201\|20 | ···················· | ···················· | ···················· | ···················· | ···················· | ···················· |
| H | 3730 | hCoV-19/England/QEUH-997D59/2020\|EPI_ISL_537202\|20 | ···················· | ···················· | ···················· | ···················· | ···················· | ···················· |
| H | 3731 | hCoV-19/England/QEUH-997E65/2020\|EPI_ISL_537204\|20 | ···················· | ···················· | ···················· | ···················· | ···················· | ···················· |
| H | 3732 | hCoV-19/England/QEUH-997D68/2020\|EPI_ISL_537205\|20 | ···················· | ···················· | ···················· | ···················· | ···················· | ···················· |
| H | 3733 | hCoV-19/England/SHEF-10B30FA/2020\|EPI_ISL_537206\|2 | ···················· | ···················· | ···················· | ···················· | ···················· | ···················· |
| H | 3734 | hCoV-19/England/SHEF-10B2A93/2020\|EPI_ISL_537208\|2 | ···················· | ···················· | ···················· | ···················· | ···················· | ···················· |
| H | 3735 | hCoV-19/England/SHEF-10B2E6A/2020\|EPI_ISL_537209\|2 | ···················· | ···················· | ···················· | ···················· | ···················· | ···················· |
| H | 3736 | hCoV-19/England/SHEF-10B2A39/2020\|EPI_ISL_537212\|2 | ···················· | ···················· | ···················· | ···················· | ···················· | ···················· |
| H | 3737 | hCoV-19/England/SHEF-10B2D5E/2020\|EPI_ISL_537214\|2 | ···················· | ···················· | ···················· | ···················· | ···················· | ···················· |
| H | 3738 | hCoV-19/England/SHEF-10B2E3D/2020\|EPI_ISL_537215\|2 | ···················· | ···················· | ···················· | ···················· | ···················· | ···················· |
| H | 3739 | hCoV-19/England/SHEF-10B2E1F/2020\|EPI_ISL_537216\|2 | ···················· | ···················· | ···················· | ···················· | ···················· | ···················· |
| H | 3740 | hCoV-19/England/SHEF-10B2E2E/2020\|EPI_ISL_537220\|2 | ···················· | ···················· | ···················· | ···················· | ···················· | ···················· |
| H | 3741 | hCoV-19/England/SHEF-10B2E4C/2020\|EPI_ISL_537221\|2 | ···················· | ···················· | ···················· | ···················· | ···················· | ···················· |
| H | 3742 | hCoV-19/England/SHEF-10B2A0C/2020\|EPI_ISL_537223\|2 | ···················· | ···················· | ···················· | ···················· | ···················· | ···················· |
| H | 3743 | hCoV-19/England/SHEF-10B2AC0/2020\|EPI_ISL_537224\|2 | ···················· | ···················· | ···················· | ···················· | ···················· | ···················· |
| H | 3744 | hCoV-19/England/SHEF-10B2D8B/2020\|EPI_ISL_537225\|2 | ···················· | ···················· | ···················· | ···················· | ···················· | ···················· |
| H | 3745 | hCoV-19/England/SHEF-10B30BE/2020\|EPI_ISL_537226\|2 | ···················· | ···················· | ···················· | ···················· | ···················· | ···················· |
| H | 3746 | hCoV-19/England/SHEF-10B2B54/2020\|EPI_ISL_537227\|2 | ···················· | ···················· | ···················· | ···················· | ···················· | ···················· |
| H | 3747 | hCoV-19/England/SHEF-10B2A48/2020\|EPI_ISL_537229\|2 | ···················· | ···················· | ···················· | ···················· | ···················· | ···················· |
| H | 3748 | hCoV-19/England/SHEF-10B2E88/2020\|EPI_ISL_537230\|2 | ···················· | ···················· | ···················· | ···················· | ···················· | ···················· |
| H | 3749 | hCoV-19/England/SHEF-10B2A66/2020\|EPI_ISL_537231\|2 | ···················· | ···················· | ···················· | ···················· | ···················· | ···················· |
| H | 3750 | hCoV-19/England/SHEF-10B2DC7/2020\|EPI_ISL_537237\|2 | ···················· | ···················· | ···················· | ···················· | ···················· | ···················· |
| H | 3751 | hCoV-19/England/SHEF-10B2D12/2020\|EPI_ISL_537238\|2 | ···················· | ···················· | ···················· | ···················· | ···················· | ···················· |
| H | 3752 | hCoV-19/England/SHEF-10B2E5B/2020\|EPI_ISL_537239\|2 | ···················· | ···················· | ···················· | ···················· | ···················· | ···················· |
| H | 3753 | hCoV-19/England/SHEF-10B2CE8/2020\|EPI_ISL_537241\|2 | ···················· | ···················· | ···················· | ···················· | ···················· | ···················· |
| H | 3754 | hCoV-19/England/SHEF-10B3090/2020\|EPI_ISL_537243\|2 | ···················· | ···················· | ···················· | ···················· | ···················· | ···················· |
| H | 3755 | hCoV-19/England/SHEF-10B2C9D/2020\|EPI_ISL_537244\|2 | ···················· | ···················· | ···················· | ···················· | ···················· | ···················· |
| H | 3756 | hCoV-19/England/SHEF-10B290F/2020\|EPI_ISL_537280\|2 | ···················· | ···················· | ···················· | ···················· | ···················· | ···················· |
| H | 3757 | hCoV-19/England/SHEF-10B285D/2020\|EPI_ISL_537282\|2 | ···················· | ···················· | ···················· | ···················· | ···················· | ···················· |
| H | 3758 | hCoV-19/England/SHEF-10B2802/2020\|EPI_ISL_537283\|2 | ···················· | ···················· | ···················· | ···················· | ···················· | ···················· |

```
H 3759 hCoV-19/England/SHEF-10B286C/2020|EPI_ISL_537284|2 ···················· ···················· ···················· ···················· ···················· ····················
H 3760 hCoV-19/Australia/NSW-SAViD-2751/2020|EPI_ISL_5372 ···················· ···················· ···················· ···················· ···················· ····················
H 3761 hCoV-19/Australia/NSW-SAViD-2752/2020|EPI_ISL_5372 ···················· ···················· ···················· ···················· ···················· ····················
H 3769 hCoV-19/Spain/CL-IBV-97006149/2020|EPI_ISL_537296| ···················· ···················· ···················· ···················· ···················· ····················
H 3773 hCoV-19/Spain/CT-IrsiCaixa-2020090134-2/2020|EPI_I ···················· ···················· ···················· ···················· ···················· ····················
H 3774 hCoV-19/USA/SC-DHEC-1196/2020|EPI_ISL_539879|2020- ···················· ···················· ···················· ···················· ···················· ····················
H 3775 hCoV-19/England/203740645/2020|EPI_ISL_539880|2020 ···················· ···················· ···················· ···················· ···················· ····················
H 3776 hCoV-19/Sweden/20-19745/2020|EPI_ISL_539881|2020-0 ···················· ···················· ···················· ···················· ···················· ····················
H 3777 hCoV-19/Sweden/20-51958/2020|EPI_ISL_539882|2020-0 ···················· ···················· ···················· ···················· ···················· ····················
H 3778 hCoV-19/Sweden/20-52156/2020|EPI_ISL_539883|2020-0 ···················· ···················· ···················· ···················· ···················· ····················
H 3779 hCoV-19/Sweden/20-52440/2020|EPI_ISL_539884|2020-0 ···················· ···················· ···················· ···················· ···················· ····················
H 3780 hCoV-19/Sweden/20-52453/2020|EPI_ISL_539885|2020-0 ···················· ···················· ···················· ···················· ···················· ····················
H 3781 hCoV-19/Sweden/20-52480/2020|EPI_ISL_539886|2020-0 ···················· ···················· ···················· ···················· ···················· ····················
H 3782 hCoV-19/England/BRIS-1334E9/2020|EPI_ISL_539894|20 ···················· ···················· ···················· ···················· ···················· ····················
H 3783 hCoV-19/England/BRIS-13355F/2020|EPI_ISL_539896|20 ···················· ···················· ···················· ···················· ···················· ····················
H 3784 hCoV-19/England/QEUH-98FB0F/2020|EPI_ISL_539897|20 ···················· ···A················ ···················· ···················· ···················· ····················
H 3785 hCoV-19/England/QEUH-98F97E/2020|EPI_ISL_539898|20 ···················· ···················· ···················· ···················· ···················· ····················
H 3786 hCoV-19/Scotland/CVR4155/2020|EPI_ISL_540840|2020- ···················· ···················· ···················· ···················· ···················· ····················
H 3787 hCoV-19/Scotland/CVR4156/2020|EPI_ISL_540841|2020- ···················· ···················· ···················· ···················· ···················· ····················
H 3788 hCoV-19/Scotland/CVR4158/2020|EPI_ISL_540842|2020- ···················· ···················· ···················· ···················· ···················· ····················
H 3789 hCoV-19/Scotland/CVR4162/2020|EPI_ISL_540843|2020- ···················· ···················· ···················· ···················· ···················· ····················
H 3790 hCoV-19/Scotland/CVR4163/2020|EPI_ISL_540844|2020- ···················· ···················· ···················· ···················· ···················· ····················
H 3791 hCoV-19/Scotland/CVR4165/2020|EPI_ISL_540846|2020- ···················· ···················· ···················· ···················· ···················· ····················
H 3792 hCoV-19/Scotland/CVR4166/2020|EPI_ISL_540847|2020- ···················· ···················· ···················· ···················· ···················· ····················
H 3793 hCoV-19/Scotland/CVR4169/2020|EPI_ISL_540849|2020- ···················· ···················· ···················· ···················· ···················· ····················
H 3794 hCoV-19/Scotland/CVR4174/2020|EPI_ISL_540851|2020- ···················· ···················· ···················· ···················· ···················· ····················
H 3795 hCoV-19/Scotland/CVR4175/2020|EPI_ISL_540852|2020- ···················· ···················· ···················· ···················· ···················· ····················
H 3796 hCoV-19/Scotland/CVR4176/2020|EPI_ISL_540853|2020- ···················· ···················· ···················· ···················· ···················· ····················
H 3797 hCoV-19/Scotland/CVR4178/2020|EPI_ISL_540855|2020- ···················· ···················· ···················· ···················· ···················· ····················
H 3798 hCoV-19/Scotland/CVR4180/2020|EPI_ISL_540856|2020- ···················· ···················· ···················· ···················· ···················· ····················
H 3799 hCoV-19/Scotland/CVR4181/2020|EPI_ISL_540857|2020- ···················· ···A················ ···················· ···················· ···················· ····················
H 3800 hCoV-19/Scotland/CVR4182/2020|EPI_ISL_540858|2020- ···················· ···················· ···················· ···················· ···················· ····················
H 3801 hCoV-19/Scotland/CVR4183/2020|EPI_ISL_540859|2020- ···················· ···················· ···················· ···················· ···················· ····················
H 3802 hCoV-19/Scotland/CVR4188/2020|EPI_ISL_540861|2020- ···················· ···················· ···················· ···················· ···················· ····················
H 3803 hCoV-19/Scotland/CVR4191/2020|EPI_ISL_540863|2020- ···················· ···················· ···················· ···················· ···················· ····················
H 3804 hCoV-19/Scotland/CVR4192/2020|EPI_ISL_540864|2020- ···················· ···················· ···················· ···················· ···················· ····················
H 3805 hCoV-19/Scotland/CVR4196/2020|EPI_ISL_540866|2020- ···················· ···················· ···················· ···················· ···················· ····················
H 3806 hCoV-19/Scotland/CVR4205/2020|EPI_ISL_540870|2020- ···················· ···················· ···················· ···················· ···················· ····················
H 3807 hCoV-19/Scotland/EDB7381/2020|EPI_ISL_540872|2020- ···················· ···················· ···················· ···················· ···················· ····················
H 3808 hCoV-19/Scotland/EDB7736/2020|EPI_ISL_540880|2020- ···················· ·A·················· ···················· ···················· ···················· ··A·················
H 3809 hCoV-19/Scotland/EDB7739/2020|EPI_ISL_540882|2020- ···················· ···················· ···················· ···················· ···················· ····················
H 3810 hCoV-19/Scotland/EDB7743/2020|EPI_ISL_540883|2020- ···················· ···················· ···················· ···················· ···················· ····················
H 3811 hCoV-19/Scotland/EDB7745/2020|EPI_ISL_540884|2020- ···················· ···················· ···················· ···················· ···················· ····················
H 3812 hCoV-19/Scotland/EDB7746/2020|EPI_ISL_540885|2020- ···················· ···················· ···················· ···················· ···················· ····················
H 3813 hCoV-19/Scotland/EDB7748/2020|EPI_ISL_540886|2020- ···················· ···················· ···················· ···················· ···················· ····················
H 3814 hCoV-19/Scotland/EDB7752/2020|EPI_ISL_540889|2020- ···················· ···················· ···················· ···················· ···················· ····················
H 3815 hCoV-19/Scotland/EDB7761/2020|EPI_ISL_540892|2020- ···················· ···················· ···················· ···················· ···················· ····················
H 3816 hCoV-19/Scotland/EDB7762/2020|EPI_ISL_540893|2020- ···················· ···················· ···················· ···················· ···················· ····················
H 3817 hCoV-19/USA/TX-HMH-3544/2020|EPI_ISL_543007|2020-0 ···················· ···················· ···················· ···················· ···················· ····················
H 3819 hCoV-19/USA/TX-HMH-3546/2020|EPI_ISL_543009|2020-0 ···················· ···················· ···················· ···················· ···················· ····················
H 3823 hCoV-19/USA/TX-HMH-3553/2020|EPI_ISL_543015|2020-0 ···················· ···················· ···················· ···················· ···················· ····················
H 3824 hCoV-19/USA/TX-HMH-3555/2020|EPI_ISL_543017|2020-0 ···················· ···················· ···················· ···················· ···················· ····················
H 3825 hCoV-19/USA/TX-HMH-3556/2020|EPI_ISL_543018|2020-0 ···················· ···················· ···················· ···················· ···················· ····················
H 3828 hCoV-19/USA/TX-HMH-3561/2020|EPI_ISL_543023|2020-0 ···················· ···················· ···················· ···················· ···················· ····················
H 3833 hCoV-19/USA/TX-HMH-3570/2020|EPI_ISL_543030|2020-0 ···················· ···················· ···················· ···················· ···················· ····················
H 3834 hCoV-19/USA/TX-HMH-3571/2020|EPI_ISL_543031|2020-0 ···················· ···················· ···················· ···················· ···················· ··A·················
H 3838 hCoV-19/USA/TX-HMH-3576/2020|EPI_ISL_543036|2020-0 ···················· ···················· ···················· ···················· ··············N····· NN····N············· 1
H 3840 hCoV-19/Nigeria/NG57839/2020|EPI_ISL_654794|2020-0 ···················· ···················· ···················· ···················· ···················· ····················
H 3841 hCoV-19/Scotland/QEUH-531247/2020|EPI_ISL_532200|2 ···················· ···················· ···················· ···················· ···················· ····················
H 3842 hCoV-19/England/QEUH-551C52/2020|EPI_ISL_532201|20 ···················· ···················· ···················· ···················· ···················· ····················
H 3843 hCoV-19/Scotland/QEUH-54EE34/2020|EPI_ISL_532202|2 ···················· ···················· ···················· ···················· ···················· ····················
H 3844 hCoV-19/Scotland/QEUH-554F56/2020|EPI_ISL_532203|2 ···················· ···················· ···················· ···················· ···················· ····················
H 3845 hCoV-19/England/QEUH-55376E/2020|EPI_ISL_532204|20 ···················· ···················· ···················· ···················· ···················· ····················
H 3846 hCoV-19/England/QEUH-555D97/2020|EPI_ISL_532205|20 ···················· ···················· ···················· ···················· ···················· ····················
H 3847 hCoV-19/England/QEUH-556C99/2020|EPI_ISL_532206|20 ···················· ···················· ···················· ···················· ···················· ····················
H 3848 hCoV-19/England/QEUH-539E96/2020|EPI_ISL_532209|20 ···················· ···················· ···················· ···················· ···················· ····················
H 3849 hCoV-19/Scotland/QEUH-5303FA/2020|EPI_ISL_532210|2 ···················· ···················· ···················· ···················· ···················· ····················
H 3850 hCoV-19/England/QEUH-538969/2020|EPI_ISL_532212|20 ···················· ···················· ···················· ···················· ···················· ····················
H 3851 hCoV-19/Scotland/QEUH-553BEA/2020|EPI_ISL_532214|2 ···················· ···················· ···················· ···················· ···················· ····················
H 3852 hCoV-19/England/QEUH-556A35/2020|EPI_ISL_532216|20 ···················· ···················· ···················· ···················· ···················· ····················
H 3853 hCoV-19/England/QEUH-5317C0/2020|EPI_ISL_532217|20 ···················· ···················· ···················· ···················· ···················· ····················
H 3854 hCoV-19/England/QEUH-538C9D/2020|EPI_ISL_532220|20 ···················· ···················· ···················· ···················· ···················· ····················
H 3855 hCoV-19/England/QEUH-5428D8/2020|EPI_ISL_532222|20 ···················· ···················· ···················· ···················· ···················· ····················
H 3856 hCoV-19/England/QEUH-544108/2020|EPI_ISL_532223|20 ···················· ···················· ···················· ···················· ···················· ····················
H 3857 hCoV-19/England/QEUH-54C6C5/2020|EPI_ISL_532224|20 ···················· ···················· ···················· ···················· ···················· ····················
H 3858 hCoV-19/England/QEUH-541A3D/2020|EPI_ISL_532226|20 ···················· ···················· ···················· ···················· ···················· ····················
H 3859 hCoV-19/England/QEUH-540C83/2020|EPI_ISL_532227|20 ···················· ···················· ···················· ···················· ···················· ····················
H 3860 hCoV-19/England/QEUH-531C67/2020|EPI_ISL_532228|20 ···················· ···················· ···················· ···················· ···················· ····················
H 3861 hCoV-19/England/QEUH-532792/2020|EPI_ISL_532230|20 ···················· ···················· ···················· ···················· ···················· ····················
H 3862 hCoV-19/England/QEUH-52FAAE/2020|EPI_ISL_532231|20 ···················· ···················· ···················· ···················· ···················· ····················
```

H 3863 hCoV-19/England/QEUH-53DA8F/2020|EPI_ISL_532233|20 ···················· ···················· ···················· ···················· ···················· ····················
H 3864 hCoV-19/England/QEUH-537F1D/2020|EPI_ISL_532234|20 ···················· ···················· ···················· ···················· ···················· ····················
H 3865 hCoV-19/England/QEUH-54136F/2020|EPI_ISL_532236|20 ···················· ···················· ···················· ···················· ···················· ····················
H 3866 hCoV-19/England/QEUH-5313AE/2020|EPI_ISL_532237|20 ···················· ···················· ···················· ···················· ···················· ····················
H 3867 hCoV-19/England/QEUH-53F2EF/2020|EPI_ISL_532238|20 ···················· ···················· ···················· ···················· ···················· ····················
H 3868 hCoV-19/England/QEUH-52DD3E/2020|EPI_ISL_532239|20 ···················· ···················· ···················· ···················· ···················· ····················
H 3869 hCoV-19/Scotland/QEUH-5456F9/2020|EPI_ISL_532241|2 ···················· ···················· ···················· ···················· ···················· ····················
H 3870 hCoV-19/England/QEUH-54C452/2020|EPI_ISL_532243|20 ···················· ···················· ···················· ···················· ···················· ····················
H 3871 hCoV-19/England/QEUH-5574FA/2020|EPI_ISL_532245|20 ···················· ···················· ···················· ···················· ···················· ····················
H 3872 hCoV-19/England/QEUH-541FF2/2020|EPI_ISL_532247|20 ···················· ···················· ···········T········ ···················· ···················· ····················
H 3873 hCoV-19/England/QEUH-53E918/2020|EPI_ISL_532248|20 ···················· ···················· ···················· ···················· ···················· ····················
H 3874 hCoV-19/England/QEUH-532765/2020|EPI_ISL_532251|20 ···················· ···················· ···················· ···················· ···················· ····················
H 3875 hCoV-19/England/QEUH-5411ED/2020|EPI_ISL_532252|20 ···················· ···················· ···················· ···················· ···················· ····················
H 3876 hCoV-19/England/QEUH-54E256/2020|EPI_ISL_532253|20 ···················· ···················· ···················· ···················· ···················· ····················
H 3877 hCoV-19/England/QEUH-537E01/2020|EPI_ISL_532254|20 ···················· ···················· ···················· ···················· ···················· ····················
H 3878 hCoV-19/England/QEUH-52E78A/2020|EPI_ISL_532255|20 ···················· ···················· ···················· ···················· ···················· ····················
H 3879 hCoV-19/England/QEUH-54E599/2020|EPI_ISL_532256|20 ···················· ···················· ···················· ···················· ···················· ····················
H 3880 hCoV-19/England/QEUH-537A1C/2020|EPI_ISL_532257|20 ···················· ···················· ···················· ···················· ···················· ····················
H 3881 hCoV-19/Scotland/QEUH-530CC2/2020|EPI_ISL_532258|2 ···················· ···················· ···················· ···················· ···················· ····················
H 3882 hCoV-19/England/QEUH-54DFC9/2020|EPI_ISL_532260|20 ···················· ···················· ···················· ···················· ···················· ····················
H 3883 hCoV-19/England/QEUH-53931F/2020|EPI_ISL_532261|20 ···················· ···················· ···················· ···················· ···················· ····················
H 3884 hCoV-19/England/QEUH-5319CA/2020|EPI_ISL_532262|20 ···················· ···················· ···················· ···················· ···················· ····················
H 3885 hCoV-19/Scotland/QEUH-530ECC/2020|EPI_ISL_532263|2 ···················· ···················· ···················· ···················· ···················· ····················
H 3886 hCoV-19/England/QEUH-53C7F2/2020|EPI_ISL_532265|20 ···················· ···················· ···················· ···················· ···················· ····················
H 3887 hCoV-19/England/QEUH-53CFAF/2020|EPI_ISL_532266|20 ···················· ···················· ···················· ···················· ···················· ····················
H 3888 hCoV-19/England/QEUH-55432D/2020|EPI_ISL_532267|20 ···················· ···················· ···················· ···················· ···················· ····················
H 3889 hCoV-19/England/QEUH-5338DA/2020|EPI_ISL_532268|20 ···················· ·A·················· ···················· ···················· ···················· ····················
H 3890 hCoV-19/England/QEUH-53EB9A/2020|EPI_ISL_532269|20 ···················· ···················· ···················· ···················· ···················· ····················
H 3891 hCoV-19/England/QEUH-551E10/2020|EPI_ISL_532270|20 ···················· ···················· ···················· ···················· ···················· ····················
H 3892 hCoV-19/Scotland/QEUH-55418D/2020|EPI_ISL_532271|2 ···················· ···················A ···················· ···················· ···················· ···················· 1
H 3893 hCoV-19/Scotland/QEUH-53A5AF/2020|EPI_ISL_532272|2 ···················· ···················A ···················· ···················· ···················· ···················· 1
H 3894 hCoV-19/England/QEUH-533861/2020|EPI_ISL_532273|20 ···················· ···················· ···················· ···················· ···················· ····················
H 3895 hCoV-19/Scotland/QEUH-530248/2020|EPI_ISL_532275|2 ···················· ···················· ···················· ···················· ···················· ····················
H 3896 hCoV-19/Scotland/QEUH-5509B6/2020|EPI_ISL_532276|2 ···················· ···················A ···················· ···················· ···················· ···················· 1
H 3897 hCoV-19/Scotland/QEUH-532D09/2020|EPI_ISL_532278|2 ···················· ···················A ···················· ···················· ···················· ···················· 1
H 3898 hCoV-19/England/QEUH-53C2C4/2020|EPI_ISL_532279|20 ···················· ···················· ···················· ···················· ···················· ····················
H 3899 hCoV-19/England/QEUH-551900/2020|EPI_ISL_532280|20 ···················· ···················· ···················· ···················· ···················· ····················
H 3900 hCoV-19/England/QEUH-5538A7/2020|EPI_ISL_532282|20 ···················· ···················· ···················· ···················· ···················· ····················
H 3901 hCoV-19/Scotland/QEUH-95F740/2020|EPI_ISL_532283|2 ···················· ···················· ···················· ···················· ···················· ····················
H 3902 hCoV-19/England/QEUH-95F081/2020|EPI_ISL_532284|20 ···················· ···················· ···················· ···················· ···················· ····················
H 3903 hCoV-19/Northern Ireland/QEUH-95EC60/2020|EPI_ISL_ ···················· ···················· ···················· ···················· ···················· ····················
H 3904 hCoV-19/England/QEUH-95EA84/2020|EPI_ISL_532286|20 ···················· ·A·················· ···················· ···················· ···················· ····················
H 3905 hCoV-19/England/QEUH-95E5FC/2020|EPI_ISL_532287|20 ···················· ···················· ···················· ···················· ···················· ····················
H 3906 hCoV-19/Scotland/QEUH-95F0DC/2020|EPI_ISL_532288|2 ···················· ···················· ···················· ···················· ···················· ····················
H 3907 hCoV-19/England/QEUH-95ED8B/2020|EPI_ISL_532289|20 ···················· ···················· ···················· ···················· ···················· ····················
H 3908 hCoV-19/England/QEUH-95F1BB/2020|EPI_ISL_532290|20 ···················· ···················· ···················· ···················· ···················· ····················
H 3909 hCoV-19/Scotland/QEUH-95E592/2020|EPI_ISL_532293|2 ···················· ···················· ···················· ···················· ···················· ····················
H 3910 hCoV-19/England/QEUH-95FB17/2020|EPI_ISL_532295|20 ···················· ···················· ···················· ···················· ···················· ····················
H 3911 hCoV-19/Northern Ireland/QEUH-95E9E1/2020|EPI_ISL_ ···················· ···················· ···················· ···················· ···················· ····················
H 3912 hCoV-19/Northern Ireland/QEUH-95EC24/2020|EPI_ISL_ ···················· ···················· ···················· ···················· ···················· ····················
H 3913 hCoV-19/Northern Ireland/QEUH-95EB54/2020|EPI_ISL_ ···················· ···················· ···················· ···················· ···················· ····················
H 3914 hCoV-19/England/QEUH-95F1E8/2020|EPI_ISL_532299|20 ···················· ·A·················· ···················· ···················· ···················· ····················
H 3915 hCoV-19/England/OXON-B31FA/2020|EPI_ISL_534844|202 ···················· ···················· ···················· ···················· ···················· ····················
H 3916 hCoV-19/England/OXON-B3215/2020|EPI_ISL_534845|202 ···················· ···················· ···················· ···················· ···················· ····················
H 3917 hCoV-19/England/OXON-B328E/2020|EPI_ISL_534846|202 ···················· ···················· ···················· ···················· ···················· ····················
H 3918 hCoV-19/England/OXON-B329D/2020|EPI_ISL_534847|202 ···················· ···················· ···················· ···················· ···················· ····················
H 3919 hCoV-19/England/OXON-B32BB/2020|EPI_ISL_534848|202 ··················T· ···················· ···················· ···················· ···················· ···················· 1
H 3920 hCoV-19/England/OXON-B32D9/2020|EPI_ISL_534849|202 ···················· ···················· ···················· ···················· ···················· ····················
H 3921 hCoV-19/England/OXON-B3312/2020|EPI_ISL_534850|202 ···················· ···················· ···················· ···················· ···················· ····················
H 3922 hCoV-19/England/OXON-B3321/2020|EPI_ISL_534851|202 ···················· ···················· ···················· ···················· ···················· ····················
H 3923 hCoV-19/England/OXON-B3330/2020|EPI_ISL_534852|202 ···················· ···················· ···················· ···················· ···················· ····················
H 3924 hCoV-19/England/OXON-B334F/2020|EPI_ISL_534853|202 ···················· ···················· ···················· ···················· ···················· ····················
H 3925 hCoV-19/England/OXON-B337C/2020|EPI_ISL_534854|202 ···················· ···················· ···················· ···················· ···················· ····················
H 3926 hCoV-19/England/OXON-B339A/2020|EPI_ISL_534855|202 ···················· ···················· ···················· ···················· ···················· ····················
H 3927 hCoV-19/England/OXON-B33B8/2020|EPI_ISL_534856|202 ···················· ···················· ···················· ···················· ···················· ····················
H 3928 hCoV-19/England/OXON-B344C/2020|EPI_ISL_534857|202 ···················· ···················· ···················· ···················· ···················· ····················
H 3929 hCoV-19/England/OXON-B345B/2020|EPI_ISL_534858|202 ···················· ···················· ···················· ···················· ···················· ····················
H 3930 hCoV-19/England/OXON-B3488/2020|EPI_ISL_534859|202 ···················· ·A·················· ···················· ···················· ···················· ····················
H 3931 hCoV-19/England/OXON-B3497/2020|EPI_ISL_534860|202 ···················· ···················· ···················· ···················· ······T············· ····················
H 3932 hCoV-19/England/OXON-B34C4/2020|EPI_ISL_534861|202 ···················· ···················· ···················· ···················· ···················· ····················
H 3933 hCoV-19/England/OXON-B34F1/2020|EPI_ISL_534862|202 ···················· ···················· ···················· ···················· ···················· ····················
H 3934 hCoV-19/England/OXON-B350D/2020|EPI_ISL_534863|202 ···················· ···················· ···················· ···················· ···················· ····················
H 3935 hCoV-19/England/OXON-B353A/2020|EPI_ISL_534865|202 ···················· ···················· ···················· ···················· ···················· ····················
H 3936 hCoV-19/England/OXON-B3558/2020|EPI_ISL_534866|202 ···················· ···················· ···················· ···················· ···················· ····················
H 3937 hCoV-19/England/OXON-B3585/2020|EPI_ISL_534867|202 ···················· ···················· ···················· ···················· ···················· ····················
H 3938 hCoV-19/England/OXON-B35A3/2020|EPI_ISL_534868|202 ···················· ···················· ···················· ···················· ···················· ····················
H 3939 hCoV-19/England/OXON-B35C1/2020|EPI_ISL_534870|202 ···················· ···················· ···················· ···················· ···················· ····················
H 3940 hCoV-19/England/OXON-B35D0/2020|EPI_ISL_534871|202 ···················· ···················· ···················· ···················· ···················· ····················
H 3941 hCoV-19/England/OXON-B35EF/2020|EPI_ISL_534872|202 ···················· ···················· ···················· ···················· ···················· ····················
H 3942 hCoV-19/England/OXON-B35FE/2020|EPI_ISL_534873|202 ···················· ···················· ···················· ···················· ···················· ····················

| | | | | | | | |
|---|---|---|---|---|---|---|---|
| H | 3943 hCoV-19/England/OXON-B3628/2020\|EPI_ISL_534874\|202 | ···················· | ···················· | ····················· | ···················· | ···················· | ···················· |
| H | 3944 hCoV-19/England/OXON-B3637/2020\|EPI_ISL_534875\|202 | ···················· | ···················· | ····················· | ···················· | ···················· | ···················· |
| H | 3945 hCoV-19/England/OXON-B3664/2020\|EPI_ISL_534876\|202 | ···················· | ···················· | ····················· | ···················· | ···················· | ···················· |
| H | 3946 hCoV-19/England/OXON-B36A0/2020\|EPI_ISL_534878\|202 | ···················· | ···················· | ····················· | ···················· | ···················· | ···················· |
| H | 3947 hCoV-19/England/OXON-B36DD/2020\|EPI_ISL_534879\|202 | ···················· | ···················· | ····················· | ···················· | ···················· | ···················· |
| H | 3948 hCoV-19/England/OXON-B36EC/2020\|EPI_ISL_534880\|202 | ···················· | ···················· | ····················· | ···················· | ···················· | ···················· |
| H | 3949 hCoV-19/England/OXON-B36FB/2020\|EPI_ISL_534881\|202 | ···················· | ···················· | ····················· | ···················· | ···················· | ···················· |
| H | 3950 hCoV-19/England/OXON-B3707/2020\|EPI_ISL_534882\|202 | ···················· | ···················· | ····················· | ···················· | ···················· | ···················· |
| H | 3951 hCoV-19/England/OXON-B3725/2020\|EPI_ISL_534883\|202 | ···················· | ···················· | ····················· | ···················· | ···················· | ···················· |
| H | 3952 hCoV-19/England/OXON-B3734/2020\|EPI_ISL_534884\|202 | ···················· | ···················· | ····················· | ···················· | ···················· | ···················· |
| H | 3953 hCoV-19/England/OXON-B3761/2020\|EPI_ISL_534885\|202 | ···················· | ···················· | ····················· | ···················· | ······T············· | ···················· |
| H | 3954 hCoV-19/England/OXON-B378F/2020\|EPI_ISL_534886\|202 | ···················· | ···················· | ····················· | ···················· | ···················· | ···················· |
| H | 3955 hCoV-19/England/OXON-B379E/2020\|EPI_ISL_534887\|202 | ···················· | ···················· | ····················· | ···················· | ···················· | ···················· |
| H | 3956 hCoV-19/England/OXON-B37AD/2020\|EPI_ISL_534888\|202 | ···················· | ···················· | ····················· | ···················· | ···················· | ···················· |
| H | 3957 hCoV-19/England/OXON-B37F8/2020\|EPI_ISL_534889\|202 | ···················· | ·············A······ | ····················· | ···················· | ···················· | ···················· |
| H | 3958 hCoV-19/England/OXON-B3813/2020\|EPI_ISL_534890\|202 | ···················· | ···················· | ····················· | ···················· | ···················· | ···················· |
| H | 3959 hCoV-19/England/OXON-B387D/2020\|EPI_ISL_534891\|202 | ···················· | ···················· | ····················· | ···················· | ···················· | ···················· |
| H | 3960 hCoV-19/England/OXON-B389B/2020\|EPI_ISL_534892\|202 | ···················· | ···················· | ····················· | ···················· | ···················· | ···················· |
| H | 3961 hCoV-19/England/OXON-B38F5/2020\|EPI_ISL_534893\|202 | ···················· | ···················· | ····················· | ···················· | ···················· | ···················· |
| H | 3962 hCoV-19/England/OXON-B392F/2020\|EPI_ISL_534894\|202 | ···················· | ···················· | ····················· | ···················· | ···················· | ···················· |
| H | 3963 hCoV-19/England/OXON-B393E/2020\|EPI_ISL_534895\|202 | ···················· | ···················· | ····················· | ···················· | ···················· | ···················· |
| H | 3964 hCoV-19/England/OXON-B39E3/2020\|EPI_ISL_534896\|202 | ···················· | ···················· | ····················· | ···················· | ···················· | ···················· |
| H | 3965 hCoV-19/England/OXON-B3A2C/2020\|EPI_ISL_534897\|202 | ···················· | ···················· | ····················· | ···················· | ···················· | ···················· |
| H | 3966 hCoV-19/England/OXON-B3A4A/2020\|EPI_ISL_534898\|202 | ···················· | ···················· | ····················· | ···················· | ···················· | ···················· |
| H | 3967 hCoV-19/England/OXON-B3A68/2020\|EPI_ISL_534899\|202 | ···················· | ···················· | ····················· | ···················· | ···················· | ···················· |
| H | 3968 hCoV-19/England/OXON-B3AC2/2020\|EPI_ISL_534900\|202 | ···················· | ···················· | ····················· | ···················· | ···················· | ···················· |
| H | 3969 hCoV-19/England/OXON-B3B29/2020\|EPI_ISL_534901\|202 | ···················· | ···················· | ····················· | ···················· | ···················· | ···················· |
| H | 3970 hCoV-19/England/OXON-B3B38/2020\|EPI_ISL_534902\|202 | ···················· | ···················· | ····················· | ···················· | ···················· | ···················· |
| H | 3971 hCoV-19/England/OXON-B3B47/2020\|EPI_ISL_534903\|202 | ···················· | ···················· | ····················· | ···················· | ···················· | ···················· |
| H | 3972 hCoV-19/England/OXON-B3B83/2020\|EPI_ISL_534904\|202 | ···················· | ···················· | ····················· | ···················· | ···················· | ···················· |
| H | 3973 hCoV-19/England/OXON-B3B92/2020\|EPI_ISL_534905\|202 | ···················· | ···················· | ····················· | ···················· | ···················· | ···················· |
| H | 3974 hCoV-19/England/OXON-B3BCF/2020\|EPI_ISL_534906\|202 | ···················· | ···················· | ····················· | ···················· | ···················· | ···················· |
| H | 3975 hCoV-19/England/OXON-B3BDE/2020\|EPI_ISL_534907\|202 | ···················· | ···················· | ····················· | ···················· | ···················· | ···················· |
| H | 3976 hCoV-19/England/OXON-B3CAE/2020\|EPI_ISL_534909\|202 | ···················· | ···················· | ····················· | ···················· | ···················· | ···················· |
| H | 3977 hCoV-19/England/QEUH-997E47/2020\|EPI_ISL_537160\|20 | ···················· | ···················· | ····················· | ···················· | ···················· | ···················· |
| H | 3978 hCoV-19/England/QEUH-997EDE/2020\|EPI_ISL_537161\|20 | ···················· | ···················· | ····················· | ···················· | ···················· | ···················· |
| H | 3979 hCoV-19/England/QEUH-99806E/2020\|EPI_ISL_537162\|20 | ···················· | ···················· | ····················· | ···················· | ···················· | ···················· |
| H | 3980 hCoV-19/England/QEUH-997EB0/2020\|EPI_ISL_537163\|20 | ···················· | ···················· | ····················· | ···················· | ···················· | ···················· |
| H | 3981 hCoV-19/England/QEUH-997F44/2020\|EPI_ISL_537164\|20 | ···················· | ···················· | ····················· | ···················· | ···················· | ···················· |
| H | 3982 hCoV-19/England/QEUH-997F53/2020\|EPI_ISL_537165\|20 | ···················· | ···················· | ····················· | ···················· | ···················· | ···················· |
| H | 3983 hCoV-19/England/QEUH-997E38/2020\|EPI_ISL_537166\|20 | ···················· | ···················· | ····················· | ···················· | ···················· | ···················· |
| H | 3984 hCoV-19/England/QEUH-997FDB/2020\|EPI_ISL_537167\|20 | ···················· | ···················· | ····················· | ···················· | ···················· | ···················· |
| H | 3985 hCoV-19/England/QEUH-9980C8/2020\|EPI_ISL_537168\|20 | ···················· | ···················· | ····················· | ···················· | ···················· | ···················· |
| H | 3986 hCoV-19/England/QEUH-997D2C/2020\|EPI_ISL_537169\|20 | ···················· | ···················· | ····················· | ···················· | ···················· | ···················· |
| H | 3987 hCoV-19/England/QEUH-997E83/2020\|EPI_ISL_537170\|20 | ···················· | ·A·················· | ····················· | ···················· | ···················· | ···················· |
| H | 3988 hCoV-19/England/QEUH-997EED/2020\|EPI_ISL_537171\|20 | ···················· | ·A·················· | ····················· | ···················· | ···················· | ···················· |
| H | 3989 hCoV-19/England/QEUH-997E1A/2020\|EPI_ISL_537172\|20 | ···················· | ···················· | ····················· | ···················· | ···················· | ···················· |
| H | 3990 hCoV-19/England/QEUH-997ECF/2020\|EPI_ISL_537173\|20 | ···················· | ···················· | ····················· | ···················· | ···················· | ···················· |
| H | 3991 hCoV-19/England/QEUH-998295/2020\|EPI_ISL_537174\|20 | ···················· | ·A·················· | ····················· | ···················· | ···················· | ···················· |
| H | 3992 hCoV-19/England/QEUH-997F62/2020\|EPI_ISL_537175\|20 | ···················· | ·A·················· | ····················· | ···················· | ···················· | ···················· |
| H | 3993 hCoV-19/England/QEUH-9980B9/2020\|EPI_ISL_537176\|20 | ···················· | ···················· | ····················· | ···················· | ···················· | ···················· |
| H | 3994 hCoV-19/England/QEUH-997F35/2020\|EPI_ISL_537177\|20 | ···················· | ···················· | ····················· | ···················· | ···················· | ···················· |
| H | 3995 hCoV-19/England/QEUH-99807D/2020\|EPI_ISL_537179\|20 | ···················· | ···················· | ····················· | ···················· | ····T··············· | ···················· |
| H | 3996 hCoV-19/England/QEUH-997F71/2020\|EPI_ISL_537180\|20 | ···················· | ·A·················· | ····················· | ···················· | ···················· | ···················· |
| H | 3997 hCoV-19/England/QEUH-9981B6/2020\|EPI_ISL_537181\|20 | ···················· | ···················· | ····················· | ···················· | ···················· | ···················· |
| H | 3998 hCoV-19/England/QEUH-99809B/2020\|EPI_ISL_537184\|20 | ···················· | ···················· | ····················· | ···················· | ···················· | ···················· |
| H | 3999 hCoV-19/England/QEUH-997E0B/2020\|EPI_ISL_537185\|20 | ···················· | ···················· | ····················· | ···················· | ···················· | ···················· |
| H | 4000 hCoV-19/England/SHEF-10B2BBE/2020\|EPI_ISL_537246\|2 | ···················· | ···················· | ····················· | ···················· | ···················· | ···················· |
| H | 4001 hCoV-19/England/SHEF-10B2E79/2020\|EPI_ISL_537248\|2 | ···················· | ···················· | ····················· | ···················· | ···················· | ···················· |
| H | 4002 hCoV-19/England/SHEF-10B2CCA/2020\|EPI_ISL_537249\|2 | ···················· | ···················· | ····················· | ···················· | ···················· | ···················· |
| H | 4003 hCoV-19/England/SHEF-10B2DD6/2020\|EPI_ISL_537251\|2 | ···················· | ···················· | ····················· | ···················· | ···················· | ···················· |
| H | 4004 hCoV-19/England/SHEF-10B3081/2020\|EPI_ISL_537252\|2 | ···················· | ···················· | ····················· | ···················· | ···················· | ···················· |
| H | 4005 hCoV-19/England/SHEF-10B2DF4/2020\|EPI_ISL_537253\|2 | ···················· | ···················· | ····················· | ···················· | ···················· | ···················· |
| H | 4006 hCoV-19/England/SHEF-10B2D9A/2020\|EPI_ISL_537255\|2 | ···················· | ···················· | ····················· | ···················· | ···················· | ···················· |
| H | 4007 hCoV-19/England/SHEF-10B2B45/2020\|EPI_ISL_537256\|2 | ···················· | ···················· | ····················· | ···················· | ···················· | ···················· |
| H | 4008 hCoV-19/England/SHEF-10B2FD0/2020\|EPI_ISL_537257\|2 | ···················· | ···················· | ····················· | ···················· | ···················· | ···················· |
| H | 4009 hCoV-19/England/SHEF-10B3151/2020\|EPI_ISL_537258\|2 | ···················· | ···················· | ····················· | ···················· | ···················· | ···················· |
| H | 4010 hCoV-19/England/SHEF-10B2FA3/2020\|EPI_ISL_537259\|2 | ···················· | ···················· | ····················· | ···················· | ···················· | ···················· |
| H | 4011 hCoV-19/England/SHEF-10B2D7C/2020\|EPI_ISL_537261\|2 | ···················· | ···················· | ····················· | ···················· | ···················· | ···················· |
| H | 4012 hCoV-19/England/SHEF-10B2CBB/2020\|EPI_ISL_537263\|2 | ···················· | ···················· | ····················· | ···················· | ···················· | ···················· |
| H | 4013 hCoV-19/England/SHEF-10B2DE5/2020\|EPI_ISL_537264\|2 | ···················· | ···················· | ····················· | ···················· | ···················· | ···················· |
| H | 4014 hCoV-19/England/SHEF-10B2DA9/2020\|EPI_ISL_537265\|2 | ···················· | ···················· | ····················· | ···················· | ···················· | ···················· |
| H | 4015 hCoV-19/England/SHEF-10B2F49/2020\|EPI_ISL_537266\|2 | ···················· | ···················· | ····················· | ···················· | ···················· | ···················· |
| H | 4016 hCoV-19/England/SHEF-10B2F0D/2020\|EPI_ISL_537267\|2 | ···················· | ···················· | ····················· | ···················· | ···················· | ···················· |
| H | 4017 hCoV-19/England/SHEF-10B2D30/2020\|EPI_ISL_537269\|2 | ···················· | ···················· | ····················· | ···················· | ···················· | ···················· |
| H | 4018 hCoV-19/England/SHEF-10B3018/2020\|EPI_ISL_537270\|2 | ···················· | ···················· | ····················· | ···················· | ···················· | ···················· |
| H | 4019 hCoV-19/England/SHEF-10B2CAC/2020\|EPI_ISL_537272\|2 | ···················· | ···················· | ····················· | ···················· | ···················· | ···················· |
| H | 4020 hCoV-19/England/SHEF-10B2A84/2020\|EPI_ISL_537273\|2 | ···················· | ···················· | ····················· | ···················· | ···················· | ···················· |
| H | 4021 hCoV-19/England/SHEF-10B2B90/2020\|EPI_ISL_537274\|2 | ···················· | ···················· | ····················· | ···················· | ···················· | ···················· |
| H | 4022 hCoV-19/England/SHEF-10B2811/2020\|EPI_ISL_537275\|2 | ···················· | ···················· | ····················· | ···················· | ···················· | ···················· |

| | | | | | | | |
|---|---|---|---|---|---|---|---|
| H | 4023 hCoV-19/England/SHEF-10B28A8/2020\|EPI_ISL_537277\|2 | ···················· | ···················· | ···················· | ···················· | ···················· | ···················· |
| H | 4024 hCoV-19/USA/SC-DHEC-0865/2020\|EPI_ISL_538400\|2020- | ···················· | ·A·················· | ···················· | ···················· | ···················· | ···················· |
| H | 4025 hCoV-19/USA/SC-DHEC-0867/2020\|EPI_ISL_538401\|2020- | ···················· | ·A·················· | ···················· | ···················· | ···················· | ···················· |
| H | 4026 hCoV-19/USA/SC-DHEC-0868/2020\|EPI_ISL_538402\|2020- | ···················· | ·A·················· | ···················· | ···················· | ···················· | ···················· |
| H | 4027 hCoV-19/USA/SC-DHEC-0871/2020\|EPI_ISL_538404\|2020- | ···················· | ·········A·········· | ···················· | ···················· | ···················· | ···················· |
| H | 4028 hCoV-19/USA/SC-DHEC-0874/2020\|EPI_ISL_538406\|2020- | ···················· | ·········A·········· | ···················· | ···················· | ···················· | ···················· |
| H | 4029 hCoV-19/USA/SC-DHEC-0875/2020\|EPI_ISL_538407\|2020- | ···················· | ···················· | ···················· | ···················· | ···················· | ···················· |
| H | 4030 hCoV-19/Singapore/956/2020\|EPI_ISL_538490\|2020-04- | ···················· | ···················· | ···················· | ···················· | ···················· | ···················· |
| H | 4031 hCoV-19/Singapore/957/2020\|EPI_ISL_538491\|2020-04- | ···················· | ···················· | ···················· | ···················· | ···················· | ···················· |
| H | 4032 hCoV-19/Singapore/958/2020\|EPI_ISL_538492\|2020-04- | ···················· | ···················· | ···················· | ···················· | ···················· | ···················· |
| H | 4033 hCoV-19/Singapore/959/2020\|EPI_ISL_538493\|2020-04- | ···················· | ···················· | ···················· | ···················· | ···················· | ···················· |
| H | 4034 hCoV-19/Singapore/794nan/2020\|EPI_ISL_538494\|2020- | ···················· | ···················· | ···················· | ···················· | ···················· | ···················· |
| H | 4035 hCoV-19/Singapore/960/2020\|EPI_ISL_538495\|2020-04- | ···················· | ···················· | ···················· | ···················· | ···················· | ···················· |
| H | 4036 hCoV-19/Singapore/961/2020\|EPI_ISL_538496\|2020-04- | ···················· | ···················· | ···················· | ···················· | ···················· | ···················· |
| H | 4037 hCoV-19/Singapore/962/2020\|EPI_ISL_538497\|2020-04- | ···················· | ···················· | ···················· | ···················· | ···················· | ···················· |
| H | 4038 hCoV-19/Indonesia/JI-NIHRD-PME0999/2020\|EPI_ISL_53 | ···················· | ···················· | ···················· | ···················· | ···················· | ···················· |
| H | 4039 hCoV-19/Indonesia/JI-NIHRD-PME2054/2020\|EPI_ISL_53 | ···················· | ···················· | ···················· | ···················· | ···················· | ···················· |
| H | 4040 hCoV-19/India/TG-CCMB-AC568/2020\|EPI_ISL_539700\|20 | ···················· | ···················· | ···················· | ···················· | ···················· | ···················· |
| H | 4041 hCoV-19/India/TG-CCMB-AB91/2020\|EPI_ISL_539701\|202 | ···················· | ···················· | ···················· | ···················· | ···················· | ···················· |
| H | 4042 hCoV-19/India/TG-CCMB-AC214/2020\|EPI_ISL_539702\|20 | ···················· | ···················· | ···················· | ···················· | ···················· | ···················· |
| H | 4043 hCoV-19/India/TG-CCMB-AC528/2020\|EPI_ISL_539703\|20 | ···················· | ···················· | ···················· | ···················· | ···················· | ···················· |
| H | 4044 hCoV-19/India/TG-CCMB-AC329/2020\|EPI_ISL_539704\|20 | ···················· | ···················· | ···················· | ···················· | ···················· | ···················· |
| H | 4045 hCoV-19/India/TG-CCMB-AB261/2020\|EPI_ISL_539705\|20 | ···················· | ···················· | ···················· | ···················· | ···················· | ···················· |
| H | 4046 hCoV-19/India/TG-CCMB-AB16/2020\|EPI_ISL_539706\|202 | ···················· | ···················· | ···················· | ···················· | ···················· | ···················· |
| H | 4047 hCoV-19/India/TG-CCMB-AB401/2020\|EPI_ISL_539707\|20 | ···················· | ···················· | ···················· | ···················· | ···················· | ···················· |
| H | 4048 hCoV-19/India/TG-CCMB-AB966/2020\|EPI_ISL_539708\|20 | ···················· | ···················· | ···················· | ···················· | ···················· | ···················· |
| H | 4049 hCoV-19/India/TG-CCMB-AB182/2020\|EPI_ISL_539709\|20 | ···················· | ···················· | ···················· | ···················· | ····T··············· | ···················· |
| H | 4050 hCoV-19/India/TG-CCMB-AB192/2020\|EPI_ISL_539710\|20 | ···················· | ···················· | ···················· | ···················· | ···················· | ···················· |
| H | 4051 hCoV-19/India/TG-CCMB-R356/2020\|EPI_ISL_539711\|202 | ···················· | ···················· | ···················· | ···················· | ···················· | ···················· |
| H | 4052 hCoV-19/India/TG-CCMB-AA222/2020\|EPI_ISL_539712\|20 | ···················· | ···················· | ···T················ | ···················· | ···················· | ···················· |
| H | 4053 hCoV-19/India/TG-CCMB-AB226/2020\|EPI_ISL_539713\|20 | ···················· | ···················· | ···T················ | ···················· | ···················· | ···················· |
| H | 4054 hCoV-19/India/TG-CCMB-AB215/2020\|EPI_ISL_539714\|20 | ···················· | ···················· | ···T················ | ···················· | ···················· | ···················· |
| H | 4055 hCoV-19/India/TG-CCMB-AB248/2020\|EPI_ISL_539715\|20 | ···················· | ···················· | ···T················ | ···················· | ···················· | ···················· |
| H | 4056 hCoV-19/India/TG-CCMB-AA311/2020\|EPI_ISL_539716\|20 | ···················· | ···················· | ···················· | ···················· | ···················· | ···················· |
| H | 4057 hCoV-19/India/TG-CCMB-AC317/2020\|EPI_ISL_539717\|20 | ···················· | ···················· | ···T················ | ···················· | ···················· | ···················· |
| H | 4058 hCoV-19/India/TG-CCMB-AB344/2020\|EPI_ISL_539718\|20 | ···················· | ···················· | ···················· | ···················· | ···················· | ···················· |
| H | 4059 hCoV-19/India/TG-CCMB-AA591/2020\|EPI_ISL_539719\|20 | ···················· | ···················· | ···················· | ···················· | ···················· | ···················· |
| H | 4060 hCoV-19/India/TG-CCMB-R296/2020\|EPI_ISL_539720\|202 | ···················· | ···················· | ···················· | ···················· | ···················· | ···················· |
| H | 4061 hCoV-19/India/TG-CCMB-AB415/2020\|EPI_ISL_539721\|20 | ···················· | ···················· | ···················· | ···················· | ···················· | ···················· |
| H | 4062 hCoV-19/India/TG-CCMB-AA871/2020\|EPI_ISL_539722\|20 | ···················· | ···················· | ···················· | ···················· | ···················· | ···················· |
| H | 4063 hCoV-19/India/TG-CCMB-AC314/2020\|EPI_ISL_539723\|20 | ···················· | ···················· | ···················· | ···················· | ···················· | ···················· |
| H | 4064 hCoV-19/India/TG-CCMB-R338/2020\|EPI_ISL_539724\|202 | ···················· | ···················· | ···················· | ···················· | ···················· | ···················· |
| H | 4065 hCoV-19/India/TG-CCMB-AA344/2020\|EPI_ISL_539725\|20 | ···················· | ···················· | ···················· | ···················· | ···················· | ···················· |
| H | 4066 hCoV-19/India/TG-CCMB-AB610/2020\|EPI_ISL_539726\|20 | ···················· | ···················· | ···················· | ···················· | ···················· | ···················· |
| H | 4067 hCoV-19/India/TG-CCMB-AC475/2020\|EPI_ISL_539727\|20 | ···················· | ···················· | ···················· | ···················· | ···················· | ···················· |
| H | 4068 hCoV-19/India/TG-CCMB-AB603/2020\|EPI_ISL_539728\|20 | ···················· | ···················· | ···················· | ···················· | ···················· | ···················· |
| H | 4069 hCoV-19/India/TG-CCMB-AB05/2020\|EPI_ISL_539729\|202 | ···················· | ···················· | ···················· | ···················· | ···················· | ···················· |
| H | 4070 hCoV-19/India/TG-CCMB-AB509/2020\|EPI_ISL_539730\|20 | ···················· | ···················· | ···················· | ···················· | ···················· | ···················· |
| H | 4071 hCoV-19/India/TG-CCMB-AA902/2020\|EPI_ISL_539731\|20 | ···················· | ···················· | ···················· | ···················· | ···················· | ···················· |
| H | 4072 hCoV-19/India/TG-CCMB-AA903/2020\|EPI_ISL_539732\|20 | ···················· | ···················· | ···················· | ···················· | ···················· | ···················· |
| H | 4073 hCoV-19/India/TG-CCMB-AA616/2020\|EPI_ISL_539733\|20 | ···················· | ···A················ | ···················· | ···················· | ···················· | ···················· |
| H | 4074 hCoV-19/India/TG-CCMB-AA713/2020\|EPI_ISL_539734\|20 | ···················· | ···A················ | ···················· | ···················· | ···················· | ···················· |
| H | 4075 hCoV-19/India/TG-CCMB-AA818/2020\|EPI_ISL_539735\|20 | ···················· | ···················· | ···················· | ···················· | ···················· | ···················· |
| H | 4076 hCoV-19/India/TG-CCMB-AA620/2020\|EPI_ISL_539736\|20 | ···················· | ···················· | ···················· | ···················· | ···················· | ···················· |
| H | 4077 hCoV-19/India/TG-CCMB-AC338/2020\|EPI_ISL_539737\|20 | ···················· | ···················· | ···················· | ···················· | ···················· | ···················· |
| H | 4078 hCoV-19/India/TG-CCMB-AA328/2020\|EPI_ISL_539738\|20 | ···················· | ···················· | ···················· | ···················· | ···················· | ···················· |
| H | 4079 hCoV-19/England/NOTT-113106/2020\|EPI_ISL_540701\|20 | ···················· | ·A·················· | ···················· | ···················· | ···················· | ···················· |
| H | 4080 hCoV-19/England/NOTT-113115/2020\|EPI_ISL_540702\|20 | ···················· | ···················· | ···················· | ···················· | ···················· | ···················· |
| H | 4081 hCoV-19/England/NOTT-113160/2020\|EPI_ISL_540704\|20 | ···················· | ·A·················· | ···················· | ···················· | ···················· | ···················· |
| H | 4082 hCoV-19/England/NOTT-11317F/2020\|EPI_ISL_540705\|20 | ···················· | ···················· | ···················· | ···················· | ···················· | ···················· |
| H | 4083 hCoV-19/England/NOTT-11318E/2020\|EPI_ISL_540706\|20 | ···················· | ···················· | ···················· | ···················· | ···················· | ···················· |
| H | 4084 hCoV-19/England/NOTT-1131CA/2020\|EPI_ISL_540708\|20 | ···················· | ···················· | ···················· | ···················· | ···················· | ···················· |
| H | 4085 hCoV-19/England/NOTT-1131E8/2020\|EPI_ISL_540710\|20 | ···················· | ·A·················· | ···················· | ···················· | ···················· | ···A················ |
| H | 4086 hCoV-19/England/NOTT-113212/2020\|EPI_ISL_540711\|20 | ···················· | ·A·················· | ···················· | ···················· | ···················· | ···················· |
| H | 4087 hCoV-19/England/NOTT-113221/2020\|EPI_ISL_540712\|20 | ···················· | ·A·················· | ···················· | ···················· | ···················· | ···················· |
| H | 4088 hCoV-19/England/NOTT-113230/2020\|EPI_ISL_540713\|20 | ···················· | ···················· | ···················· | ···················· | ···················· | ···················· |
| H | 4089 hCoV-19/England/NOTT-11325E/2020\|EPI_ISL_540715\|20 | ···················· | ···················· | ···················· | ···················· | ···················· | ···················· |
| H | 4090 hCoV-19/England/NOTT-11328B/2020\|EPI_ISL_540717\|20 | ···················· | ·A·················· | ···················· | ···················· | ···················· | ···················· |
| H | 4091 hCoV-19/England/NOTT-1132A9/2020\|EPI_ISL_540718\|20 | ···················· | ·A·················· | ···················· | ···················· | ···················· | ···A················ |
| H | 4092 hCoV-19/England/SHEF-C0B65/2020\|EPI_ISL_540719\|202 | ···················· | ···················· | ···················· | ···················· | ···················· | ···················· |
| H | 4093 hCoV-19/Wales/MILK-992019/2020\|EPI_ISL_540894\|2020 | ···················· | ···················· | ···················· | ···················· | ···················· | ···················· |
| H | 4094 hCoV-19/Wales/MILK-992028/2020\|EPI_ISL_540895\|2020 | ···················· | ···················· | ···················· | ···················· | ···················· | ···················· |
| H | 4095 hCoV-19/Wales/PHWC-168524/2020\|EPI_ISL_540898\|2020 | ···················· | ···················· | ···················· | ···················· | ···················· | ···················· |
| H | 4096 hCoV-19/Wales/PHWC-16A15B/2020\|EPI_ISL_540899\|2020 | ···················· | ···················· | ···················· | ···················· | ···················· | ···················· |
| H | 4097 hCoV-19/Northern Ireland/QEUH-95ECBB/2020\|EPI_ISL_ | ···················· | ···················· | ···················· | ···················· | ···················· | ···················· |
| H | 4098 hCoV-19/Scotland/QEUH-95F1D9/2020\|EPI_ISL_532301\|2 | ···················· | ···················· | ···················· | ···················· | ···················· | ···················· |
| H | 4099 hCoV-19/England/QEUH-95F5DD/2020\|EPI_ISL_532302\|20 | ···················· | ·A·················· | ···················· | ···················· | ···················· | ···················· |
| H | 4100 hCoV-19/England/OXON-B3D50/2020\|EPI_ISL_534910\|202 | ···················· | ············A······· | ···················· | ···················· | ···················· | ···················· |
| H | 4101 hCoV-19/England/OXON-B3D7E/2020\|EPI_ISL_534911\|202 | ···················· | ···················· | ···················· | ···················· | ···················· | ···················· |
| H | 4102 hCoV-19/England/OXON-B3D8D/2020\|EPI_ISL_534912\|202 | ···················· | ···················· | ···················· | ···················· | ···················· | ···················· |

```
H   4103 hCoV-19/England/OXON-B3DF6/2020|EPI_ISL_534913|202   ····················  ····················  ····················  ····················  ····················  ····················
H   4104 hCoV-19/England/OXON-B3E8A/2020|EPI_ISL_534914|202   ····················  ····················  ····················  ····················  ····················  ····················
H   4105 hCoV-19/England/OXON-B3EA8/2020|EPI_ISL_534915|202   ····················  ····················  ····················  ····················  ····················  ····················
H   4106 hCoV-19/England/OXON-B3EE4/2020|EPI_ISL_534917|202   ····················  ····················  ····················  ····················  ····················  ····················
H   4107 hCoV-19/England/OXON-B3F2D/2020|EPI_ISL_534918|202   ····················  ····················  ····················  ····················  ····················  ····················
H   4108 hCoV-19/England/OXON-B3F4B/2020|EPI_ISL_534920|202   ····················  ····················  ····················  ····················  ····················  ····················
H   4109 hCoV-19/England/OXON-B3FC3/2020|EPI_ISL_534921|202   ····················  ····················  ····················  ····················  ····················  ····················
H   4110 hCoV-19/England/OXON-B3FE1/2020|EPI_ISL_534922|202   ····················  ····················  ····················  ····················  ····················  ····················
H   4111 hCoV-19/England/OXON-B401A/2020|EPI_ISL_534924|202   ····················  ····················  ····················  ····················  ····················  ····················
H   4112 hCoV-19/England/OXON-B4038/2020|EPI_ISL_534925|202   ····················  ····················  ····················  ····················  ····················  ····················
H   4113 hCoV-19/England/OXON-B4065/2020|EPI_ISL_534926|202   ····················  ····················  ····················  ····················  ····················  ····················
H   4114 hCoV-19/England/OXON-B4083/2020|EPI_ISL_534927|202   ····················  ····················  ····················  ····················  ····················  ····················
H   4115 hCoV-19/England/OXON-B4092/2020|EPI_ISL_534928|202   ····················  ····················  ····················  ····················  ····················  ····················
H   4116 hCoV-19/England/OXON-B40A1/2020|EPI_ISL_534929|202   ····················  ····················  ····················  ····················  ····················  ····················
H   4117 hCoV-19/England/OXON-B40FC/2020|EPI_ISL_534930|202   ····················  ····················  ····················  ····················  ····················  ····················
H   4118 hCoV-19/England/OXON-B4135/2020|EPI_ISL_534931|202   ····················  ····················  ····················  ····················  ····················  ····················
H   4119 hCoV-19/England/OXON-B4144/2020|EPI_ISL_534932|202   ····················  ····················  ····················  ····················  ····················  ····················
H   4120 hCoV-19/England/OXON-B4153/2020|EPI_ISL_534933|202   ····················  ····················  ····················  ····················  ····················  ····················
H   4121 hCoV-19/England/OXON-B4162/2020|EPI_ISL_534934|202   ····················  ····················  ····················  ····················  ····················  ····················
H   4122 hCoV-19/England/OXON-B4180/2020|EPI_ISL_534935|202   ····················  ····················  ····················  ····················  ····················  ····················
H   4123 hCoV-19/England/OXON-B41F9/2020|EPI_ISL_534938|202   ····················  ····················  ····················  ····················  ····················  ····················
H   4124 hCoV-19/England/OXON-B4205/2020|EPI_ISL_534939|202   ····················  ····················  ····················  ····················  ····················  ····················
H   4125 hCoV-19/England/OXON-B4232/2020|EPI_ISL_534941|202   ····················  ····················  ····················  ····················  ····················  ····················
H   4126 hCoV-19/England/OXON-B4250/2020|EPI_ISL_534942|202   ····················  ····················  ····················  ····················  ····················  ····················
H   4127 hCoV-19/England/OXON-B428D/2020|EPI_ISL_534943|202   ····················  ····················  ····················  ····················  ····················  ····················
H   4128 hCoV-19/England/OXON-B42AB/2020|EPI_ISL_534944|202   ····················  ····················  ····················  ····················  ····················  ····················
H   4129 hCoV-19/England/OXON-B42BA/2020|EPI_ISL_534945|202   ····················  ····················  ····················  ····················  ····················  ····················
H   4130 hCoV-19/England/OXON-B434E/2020|EPI_ISL_534947|202   ····················  ····················  ····················  ····················  ····················  ····················
H   4131 hCoV-19/England/OXON-B435D/2020|EPI_ISL_534948|202   ····················  ····················  ····················  ····················  ····················  ····················
H   4132 hCoV-19/England/OXON-B43A8/2020|EPI_ISL_534950|202   ····················  ····················  ····················  ····················  ····················  ····················
H   4133 hCoV-19/England/OXON-B43F3/2020|EPI_ISL_534952|202   ····················  ····················  ····················  ····················  ····················  ····················
H   4134 hCoV-19/England/OXON-B440F/2020|EPI_ISL_534953|202   ····················  ····················  ····················  ····················  ····················  ····················
H   4135 hCoV-19/England/OXON-B442D/2020|EPI_ISL_534954|202   ····················  ····················  ····················  ····················  ····················  ····················
H   4136 hCoV-19/England/OXON-B443C/2020|EPI_ISL_534955|202   ····················  ····················  ····················  ····················  ····················  ····················
H   4137 hCoV-19/England/OXON-B44F0/2020|EPI_ISL_534958|202   ····················  ····················  ····················  ····················  ····················  ····················
H   4138 hCoV-19/England/OXON-B4539/2020|EPI_ISL_534959|202   ····················  ····················  ····················  ····················  ····················  ····················
H   4139 hCoV-19/England/OXON-B4548/2020|EPI_ISL_534960|202   ····················  ····················  ····················  ····················  ····················  ····················
H   4140 hCoV-19/England/OXON-B4557/2020|EPI_ISL_534961|202   ····················  ····················  ····················  ····················  ····················  ····················
H   4141 hCoV-19/England/OXON-B45DF/2020|EPI_ISL_534963|202   ····················  ····················  ····················  ····················  ····················  ····················
H   4142 hCoV-19/England/OXON-B4654/2020|EPI_ISL_534964|202   ····················  ····················  ····················  ····················  ····················  ····················
H   4143 hCoV-19/England/QEUH-961602/2020|EPI_ISL_537100|20   ····················  ····················  ····················  ····················  ····················  ····················
H   4144 hCoV-19/England/QEUH-95DD22/2020|EPI_ISL_537101|20   ····················  ····················  ····················  ····················  ····················  ····················
H   4145 hCoV-19/Northern Ireland/QEUH-961A33/2020|EPI_ISL_   ····················  ····················  ····················  ····················  ····················  ····················
H   4146 hCoV-19/England/QEUH-961990/2020|EPI_ISL_537103|20   ····················  ····················  ····················  ····················  ····················  ····················
H   4147 hCoV-19/England/QEUH-961D85/2020|EPI_ISL_537104|20   ····················  ·A··················  ····················  ····················  ····················  ····················
H   4148 hCoV-19/England/QEUH-961CA6/2020|EPI_ISL_537105|20   ····················  ····················  ····················  ····················  ····················  ····················
H   4149 hCoV-19/England/QEUH-9614AE/2020|EPI_ISL_537106|20   ····················  ····················  ····················  ····················  ····················  ····················
H   4150 hCoV-19/England/QEUH-961C79/2020|EPI_ISL_537107|20   ····················  ····················  ····················  ····················  ····················  ····················
H   4151 hCoV-19/England/QEUH-961C3D/2020|EPI_ISL_537108|20   ····················  ····················  ····················  ····················  ····················  ····················
H   4152 hCoV-19/England/QEUH-96158D/2020|EPI_ISL_537109|20   ····················  ····················  ····················  ····················  ····················  ····················
H   4153 hCoV-19/England/QEUH-96173C/2020|EPI_ISL_537110|20   ····················  ····················  ····················  ····················  ····················  ····················
H   4154 hCoV-19/England/QEUH-95E50B/2020|EPI_ISL_537111|20   ····················  ····················  ····················  ····················  ····················  ····················
H   4155 hCoV-19/England/QEUH-9616D5/2020|EPI_ISL_537112|20   ····················  ····················  ····················  ····················  ····················  ····················
H   4156 hCoV-19/England/QEUH-961523/2020|EPI_ISL_537113|20   ····················  ····················  ····················  ····················  ····················  ····················
H   4157 hCoV-19/England/QEUH-961936/2020|EPI_ISL_537115|20   ····················  ····················  ····················  ····················  ····················  ····················
H   4158 hCoV-19/England/QEUH-961611/2020|EPI_ISL_537116|20   ····················  ····················  ····················  ····················  ····················  ····················
H   4159 hCoV-19/England/QEUH-961B12/2020|EPI_ISL_537117|20   ····················  ····················  ····················  ····················  ····················  ····················
H   4160 hCoV-19/Northern Ireland/QEUH-961C88/2020|EPI_ISL_   ····················  ····················  ····················  ····················  ····················  ····················
H   4161 hCoV-19/England/QEUH-9617F0/2020|EPI_ISL_537119|20   ····················  ····················  ····················  ····················  ····················  ····················
H   4162 hCoV-19/England/QEUH-9616F3/2020|EPI_ISL_537120|20   ····················  ····················  ····················  ····················  ····················  ····················
H   4163 hCoV-19/England/QEUH-9618A2/2020|EPI_ISL_537121|20   ····················  ····················  ····················  ····················  ····················  ····················
H   4164 hCoV-19/England/QEUH-961DA3/2020|EPI_ISL_537123|20   ····················  ·A··················  ····················  ····················  ····················  ····················
H   4165 hCoV-19/England/QEUH-95E33E/2020|EPI_ISL_537124|20   ····················  ····················  ····················  ····················  ····················  ····················
H   4166 hCoV-19/England/QEUH-961C00/2020|EPI_ISL_537126|20   ····················  ····················  ····················  ····················  ····················  ····················
H   4167 hCoV-19/England/QEUH-96181B/2020|EPI_ISL_537127|20   ····················  ····················  ····················  ····················  ····················  ····················
H   4168 hCoV-19/England/QEUH-95DFFF/2020|EPI_ISL_537128|20   ····················  ····················  ····················  ····················  ····················  ····················
H   4169 hCoV-19/England/QEUH-9617E1/2020|EPI_ISL_537129|20   ····················  ····················  ····················  ····················  ····················  ····················
H   4170 hCoV-19/England/QEUH-961471/2020|EPI_ISL_537130|20   ····················  ····················  ····················  ····················  ····················  ····················
H   4171 hCoV-19/England/QEUH-95DEB6/2020|EPI_ISL_537131|20   ····················  ····················  ····················  ····················  ····················  ····················
H   4172 hCoV-19/England/QEUH-961DB2/2020|EPI_ISL_537132|20   ····················  ····················  ····················  ····················  ····················  ····················
H   4173 hCoV-19/England/QEUH-9615AB/2020|EPI_ISL_537133|20   ····················  ·A··················  ····················  ····················  ····················  ····················
H   4174 hCoV-19/England/QEUH-961918/2020|EPI_ISL_537134|20   ····················  ····················  ····················  ····················  ····················  ····················
H   4175 hCoV-19/Northern Ireland/QEUH-961AD9/2020|EPI_ISL_   ····················  ····················  ····················  ····················  ····················  ····················
H   4176 hCoV-19/England/QEUH-9616E4/2020|EPI_ISL_537137|20   ····················  ····················  ····················  ····················  ····················  ····················
H   4177 hCoV-19/England/QEUH-95DCE9/2020|EPI_ISL_537138|20   ····················  ····················  ····················  ····················  ····················  ····················
H   4178 hCoV-19/England/QEUH-95DE10/2020|EPI_ISL_537139|20   ····················  ····················  ····················  ····················  ····················  ····················
H   4179 hCoV-19/England/QEUH-95DDC8/2020|EPI_ISL_537140|20   ····················  ····················  ····················  ····················  ····················  ····················
H   4180 hCoV-19/England/QEUH-95DF1D/2020|EPI_ISL_537141|20   ····················  ····················  ····················  ····················  ····················  ····················
H   4181 hCoV-19/England/QEUH-961B4F/2020|EPI_ISL_537142|20   ····················  ····················  ····················  ····················  ····················  ····················
H   4182 hCoV-19/Northern Ireland/QEUH-961BF4/2020|EPI_ISL_   ····················  ····················  ····················  ····················  ····················  ····················
```

| | | | | | | | | |
|---|---|---|---|---|---|---|---|---|
| H | 4183 | hCoV-19/England/QEUH-95E055/2020\|EPI_ISL_537144\|20 | ···················· | ···················· | ···················· | ···················· | ···················· | ···················· |
| H | 4184 | hCoV-19/England/QEUH-95E36B/2020\|EPI_ISL_537145\|20 | ···················· | ·A·················· | ···················· | ···················· | ···················· | ···A················ |
| H | 4185 | hCoV-19/England/QEUH-997DC2/2020\|EPI_ISL_537149\|20 | ···················· | ···················· | ···················· | ···················· | ···················· | ···················· |
| H | 4186 | hCoV-19/England/QEUH-9980AA/2020\|EPI_ISL_537182\|20 | ···················· | ···················· | ···················· | ···················· | ···················· | ···················· |
| H | 4187 | hCoV-19/England/QEUH-99805F/2020\|EPI_ISL_537188\|20 | ···················· | ···················· | ···················· | ···················· | ···················· | ···················· |
| H | 4188 | hCoV-19/England/QEUH-998277/2020\|EPI_ISL_537189\|20 | ···················· | ···················· | ···················· | ···················· | ···················· | ···················· |
| H | 4189 | hCoV-19/England/QEUH-9981D4/2020\|EPI_ISL_537190\|20 | ···················· | ···················· | ···················· | ···················· | ···················· | ···················· |
| H | 4190 | hCoV-19/England/QEUH-997E29/2020\|EPI_ISL_537191\|20 | ···················· | ···················· | ···················· | ···················· | ···················· | ···················· |
| H | 4191 | hCoV-19/England/QEUH-9982A4/2020\|EPI_ISL_537192\|20 | ···················· | ···················· | ···················· | ···················· | ···················· | ···················· |
| H | 4192 | hCoV-19/England/QEUH-997E74/2020\|EPI_ISL_537193\|20 | ···················· | ···················· | ···················· | ···················· | ···················· | ···················· |
| H | 4193 | hCoV-19/England/QEUH-997DA4/2020\|EPI_ISL_537195\|20 | ···················· | ···················· | ···················· | ···················· | ···················· | ···················· |
| H | 4194 | hCoV-19/England/QEUH-997DB3/2020\|EPI_ISL_537196\|20 | ···················· | ···················· | ···················· | ···················· | ···················· | ···················· |
| H | 4195 | hCoV-19/England/QEUH-997F08/2020\|EPI_ISL_537198\|20 | ···················· | ···················· | ···················· | ···················· | ···················· | ···················· |
| H | 4196 | hCoV-19/England/QEUH-9981E3/2020\|EPI_ISL_537199\|20 | ···················· | ···················· | ···················· | ···················· | ···················· | ···················· |
| H | 4197 | hCoV-19/USA/SC-DHEC-0886/2020\|EPI_ISL_538410\|2020- | ···················· | ···················· | ···················· | ···················· | ···················· | ···················· |
| H | 4198 | hCoV-19/USA/SC-DHEC-0887/2020\|EPI_ISL_538411\|2020- | ···················· | ···················· | ···················· | ···················· | ···················· | ···················· |
| H | 4199 | hCoV-19/USA/SC-DHEC-0891/2020\|EPI_ISL_538412\|2020- | ···················· | ···················· | ···················· | ···················· | ···················· | ···················· |
| H | 4200 | hCoV-19/USA/SC-DHEC-0893/2020\|EPI_ISL_538413\|2020- | ···················· | ···················· | ···················· | ···················· | ···················· | ···················· |
| H | 4201 | hCoV-19/USA/SC-DHEC-0918/2020\|EPI_ISL_538415\|2020- | ···················· | ···················· | ···················· | ···················· | ···················· | ···················· |
| H | 4202 | hCoV-19/USA/SC-DHEC-0922/2020\|EPI_ISL_538417\|2020- | ···················· | ···················· | ···················· | ···················· | ···················· | ···················· |
| H | 4203 | hCoV-19/USA/SC-DHEC-0925/2020\|EPI_ISL_538418\|2020- | ···················· | ···················· | ···················· | ···················· | ···················· | ···················· |
| H | 4204 | hCoV-19/USA/SC-DHEC-0929/2020\|EPI_ISL_538420\|2020- | ···················· | ···················· | ···················· | ···················· | ···················· | ···················· |
| H | 4205 | hCoV-19/USA/SC-DHEC-0930/2020\|EPI_ISL_538421\|2020- | ···················· | ···················· | ···················· | ···················· | ···················· | ···················· |
| H | 4206 | hCoV-19/USA/SC-DHEC-0597/2020\|EPI_ISL_538422\|2020- | ···················· | ···················· | ···················· | ···················· | ···················· | ···················· |
| H | 4207 | hCoV-19/USA/SC-DHEC-0598/2020\|EPI_ISL_538423\|2020- | ···················· | ···················· | ···················· | ···················· | ···················· | ···················· |
| H | 4208 | hCoV-19/USA/SC-DHEC-0601/2020\|EPI_ISL_538425\|2020- | ···················· | ···················· | ···················· | ···················· | ···················· | ···················· |
| H | 4209 | hCoV-19/USA/SC-DHEC-0618/2020\|EPI_ISL_538426\|2020- | ···················· | ···················· | ···················· | ···················· | ···················· | ···················· |
| H | 4210 | hCoV-19/USA/SC-DHEC-0619/2020\|EPI_ISL_538427\|2020- | ···················· | ···················· | ···················· | ···················· | ···················· | ···················· |
| H | 4211 | hCoV-19/USA/SC-DHEC-0644/2020\|EPI_ISL_538428\|2020- | ···················· | ···················· | ···················· | ···················· | ···················· | ···················· |
| H | 4212 | hCoV-19/USA/SC-DHEC-0648/2020\|EPI_ISL_538430\|2020- | ···················· | ···················· | ···················· | ···················· | ···················· | ···················· |
| H | 4213 | hCoV-19/USA/SC-DHEC-0650/2020\|EPI_ISL_538431\|2020- | ···················· | ···················· | ···················· | ···················· | ···················· | ···················· |
| H | 4214 | hCoV-19/USA/SC-DHEC-0881/2020\|EPI_ISL_538432\|2020- | ···················· | ···················· | ···················· | ···················· | ···················· | ···················· |
| H | 4215 | hCoV-19/USA/SC-DHEC-0882/2020\|EPI_ISL_538433\|2020- | ···················· | ·A·················· | ···················· | ···················· | ···················· | ···················· |
| H | 4216 | hCoV-19/USA/SC-DHEC-0883/2020\|EPI_ISL_538434\|2020- | ···················· | ·A·················· | ···················· | ···················· | ···················· | ···················· |
| H | 4217 | hCoV-19/Singapore/36nan/2020\|EPI_ISL_538435\|2020-0 | ···················· | ···················· | ···················· | ···················· | ···················· | ···················· |
| H | 4218 | hCoV-19/Singapore/913/2020\|EPI_ISL_538436\|2020-02- | ···················· | ···················· | ···················· | ···················· | ···················· | ···················· |
| H | 4219 | hCoV-19/Singapore/46nan/2020\|EPI_ISL_538437\|2020-0 | ···················· | ···················· | ···················· | ···················· | ···················· | ···················· |
| H | 4220 | hCoV-19/Singapore/914/2020\|EPI_ISL_538438\|2020-03- | ···················· | ···················· | ···················· | ···················· | ···················· | ···················· |
| H | 4221 | hCoV-19/Singapore/915/2020\|EPI_ISL_538439\|2020-03- | ···················· | ···················· | ···················· | ···················· | ···················· | ···················· |
| H | 4222 | hCoV-19/Singapore/135nan/2020\|EPI_ISL_538440\|2020- | ···················· | ···················· | ···················· | ···················· | ···················· | ···················· |
| H | 4223 | hCoV-19/Singapore/173nan/2020\|EPI_ISL_538441\|2020- | ···················· | ···················· | ···················· | ···················· | ···················· | ···················· |
| H | 4224 | hCoV-19/Singapore/427nan/2020\|EPI_ISL_538442\|2020- | ···················· | ···················· | ···················· | ···················· | ···················· | ···················· |
| H | 4225 | hCoV-19/Singapore/916/2020\|EPI_ISL_538443\|2020-03- | ···················· | ···················· | ···················· | ···················· | ···················· | ···················· |
| H | 4226 | hCoV-19/Singapore/917/2020\|EPI_ISL_538444\|2020-03- | ···················· | ···················· | ···················· | ···················· | ···················· | ···················· |
| H | 4227 | hCoV-19/Singapore/918/2020\|EPI_ISL_538445\|2020-04- | ···················· | ···················· | ···················· | ···················· | ···················· | ···················· |
| H | 4228 | hCoV-19/Singapore/50nan/2020\|EPI_ISL_538446\|2020-0 | ···················· | ···················· | ···················· | ···················· | ···················· | ···················· |
| H | 4229 | hCoV-19/Singapore/148nan/2020\|EPI_ISL_538447\|2020- | ···················· | ···················· | ···················· | ···················· | ···················· | ···················· |
| H | 4230 | hCoV-19/Singapore/113nan/2020\|EPI_ISL_538448\|2020- | ···················· | ···················· | ···················· | ···················· | ···················· | ···················· |
| H | 4231 | hCoV-19/Singapore/129nan/2020\|EPI_ISL_538449\|2020- | ···················· | ···················· | ···················· | ···················· | ···················· | ···················· |
| H | 4232 | hCoV-19/Singapore/919/2020\|EPI_ISL_538450\|2020-03- | ···················· | ···················· | ···················· | ···················· | ···················· | ···················· |
| H | 4233 | hCoV-19/Singapore/920/2020\|EPI_ISL_538451\|2020-03- | ···················· | ···················· | ···················· | ···················· | ···················· | ···················· |
| H | 4234 | hCoV-19/Singapore/921/2020\|EPI_ISL_538452\|2020-03- | ···················· | ···················· | ···················· | ···················· | ···················· | ···················· |
| H | 4235 | hCoV-19/Singapore/922/2020\|EPI_ISL_538453\|2020-03- | ···················· | ···················· | ···················· | ···················· | ···················· | ···················· |
| H | 4236 | hCoV-19/Singapore/923/2020\|EPI_ISL_538454\|2020-03- | ···················· | ···················· | ···················· | ···················· | ···················· | ···················· |
| H | 4237 | hCoV-19/Singapore/924/2020\|EPI_ISL_538455\|2020-03- | ···················· | ···················· | ···················· | ···················· | ···················· | ···················· |
| H | 4238 | hCoV-19/Singapore/925/2020\|EPI_ISL_538456\|2020-03- | ···················· | ···················· | ···················· | ···················· | ···················· | ···················· |
| H | 4239 | hCoV-19/Singapore/323nan/2020\|EPI_ISL_538457\|2020- | ···················· | ···················· | ···················· | ···················· | ···················· | ···················· |
| H | 4240 | hCoV-19/Singapore/926/2020\|EPI_ISL_538458\|2020-03- | ···················· | ···················· | ···················· | ···················· | ···················· | ···················· |
| H | 4241 | hCoV-19/Singapore/927/2020\|EPI_ISL_538459\|2020-03- | ···················· | ···················· | ···················· | ···················· | ···················· | ···················· |
| H | 4242 | hCoV-19/Singapore/928/2020\|EPI_ISL_538460\|2020-03- | ···················· | ···················· | ···················· | ···················· | ···················· | ···················· |
| H | 4243 | hCoV-19/Singapore/929/2020\|EPI_ISL_538461\|2020-03- | ···················· | ···················· | ···················· | ···················· | ···················· | ···················· |
| H | 4244 | hCoV-19/Singapore/930/2020\|EPI_ISL_538462\|2020-03- | ···················· | ···················· | ···················· | ···················· | ···················· | ···················· |
| H | 4245 | hCoV-19/Singapore/931/2020\|EPI_ISL_538463\|2020-03- | ···················· | ···················· | ···················· | ···················· | ···················· | ···················· |
| H | 4246 | hCoV-19/Singapore/932/2020\|EPI_ISL_538464\|2020-03- | ···················· | ···················· | ···················· | ···················· | ···················· | ···················· |
| H | 4247 | hCoV-19/Singapore/933/2020\|EPI_ISL_538465\|2020-03- | ···················· | ···················· | ···················· | ···················· | ···················· | ···················· |
| H | 4248 | hCoV-19/Singapore/934/2020\|EPI_ISL_538466\|2020-03- | ···················· | ···················· | ···················· | ···················· | ···················· | ···················· |
| H | 4249 | hCoV-19/Singapore/217nan/2020\|EPI_ISL_538467\|2020- | ···················· | ···················· | ···················· | ···················· | ···················· | ···················· |
| H | 4250 | hCoV-19/Singapore/935/2020\|EPI_ISL_538468\|2020-04- | ···················· | ···················· | ···················· | ···················· | ···················· | ···················· |
| H | 4251 | hCoV-19/Singapore/936/2020\|EPI_ISL_538469\|2020-04- | ···················· | ···················· | ···················· | ···················· | ···················· | ···················· |
| H | 4252 | hCoV-19/Singapore/937/2020\|EPI_ISL_538470\|2020-04- | ···················· | ···················· | ···················· | ···················· | ···················· | ···················· |
| H | 4253 | hCoV-19/Singapore/564nan/2020\|EPI_ISL_538471\|2020- | ···················· | ···················· | ···················· | ···················· | ···················· | ···················· |
| H | 4254 | hCoV-19/Singapore/938/2020\|EPI_ISL_538472\|2020-04- | ···················· | ···················· | ···················· | ···················· | ···················· | ···················· |
| H | 4255 | hCoV-19/Singapore/939/2020\|EPI_ISL_538473\|2020-04- | ···················· | ···················· | ···················· | ···················· | ···················· | ···················· |
| H | 4256 | hCoV-19/Singapore/940/2020\|EPI_ISL_538474\|2020-04- | ···················· | ···················· | ···················· | ···················· | ···················· | ···················· |
| H | 4257 | hCoV-19/Singapore/941/2020\|EPI_ISL_538475\|2020-04- | ···················· | ···················· | ···················· | ···················· | ···················· | ···················· |
| H | 4258 | hCoV-19/Singapore/942/2020\|EPI_ISL_538476\|2020-04- | ···················· | ···················· | ···················· | ···················· | ···················· | ···················· |
| H | 4259 | hCoV-19/India/TG-CCMB-AA41/2020\|EPI_ISL_539739\|202 | ···················· | ·A·················· | ···················· | ···················· | ···················· | ···A················ |
| H | 4260 | hCoV-19/India/TG-CCMB-AB227/2020\|EPI_ISL_539740\|20 | ···················· | ·A·················· | ···················· | ···················· | ···················· | ···A················ |
| H | 4261 | hCoV-19/India/TG-CCMB-AC391/2020\|EPI_ISL_539741\|20 | ···················· | ·A·················· | ···················· | ···················· | ···················· | ···A················ |
| H | 4262 | hCoV-19/India/TG-CCMB-AB955/2020\|EPI_ISL_539742\|20 | ···················· | ·A·················· | ···················· | ···················· | ···················· | ···A················ |

| | | | | | | | | |
|---|---|---|---|---|---|---|---|---|
| H | 4263 | hCoV-19/India/TG-CCMB-AC222/2020\|EPI_ISL_539743\|20 | ···················· | ·A·················· | ···················· | ···················· | ···················· | ··A················· |
| H | 4264 | hCoV-19/India/TG-CCMB-AC511/2020\|EPI_ISL_539744\|20 | ···················· | ·A·················· | ···················· | ···················· | ···················· | ··A················· |
| H | 4265 | hCoV-19/India/TG-CCMB-R286/2020\|EPI_ISL_539745\|202 | ···················· | ···················· | ···················· | ···················· | ···················· | ···················· |
| H | 4266 | hCoV-19/India/TG-CCMB-AB198/2020\|EPI_ISL_539746\|20 | ···················· | ···················· | ···················· | ···················· | ···················· | ···················· |
| H | 4267 | hCoV-19/India/TG-CCMB-AA812/2020\|EPI_ISL_539747\|20 | ···················· | ···················· | ···················· | ···················· | ···················· | ···················· |
| H | 4268 | hCoV-19/India/TG-CCMB-AC512/2020\|EPI_ISL_539748\|20 | ···················· | ···················· | ···················· | ···················· | ···················· | ···················· |
| H | 4269 | hCoV-19/India/TG-CCMB-AA340/2020\|EPI_ISL_539749\|20 | ···················· | ···················· | ···················· | ···················· | ···················· | ···················· |
| H | 4270 | hCoV-19/India/TG-CCMB-AA590/2020\|EPI_ISL_539750\|20 | ···················· | ···················· | ···················· | ···················· | ···················· | ···················· |
| H | 4271 | hCoV-19/India/TG-CCMB-AA241/2020\|EPI_ISL_539751\|20 | ···················· | ···················· | ···················· | ···················· | ···················· | ···················· |
| H | 4272 | hCoV-19/India/TG-CCMB-AA272/2020\|EPI_ISL_539752\|20 | ···················· | ···················· | ···················· | ···················· | ···················· | ···················· |
| H | 4273 | hCoV-19/India/TG-CCMB-AA29/2020\|EPI_ISL_539753\|202 | ···················· | ···················· | ···················· | ···················· | ···················· | ···················· |
| H | 4274 | hCoV-19/India/TG-CCMB-AA315/2020\|EPI_ISL_539754\|20 | ···················· | ···················· | ···················· | ···················· | ···················· | ···················· |
| H | 4275 | hCoV-19/India/TG-CCMB-AA32/2020\|EPI_ISL_539755\|202 | ···················· | ···················· | ···················· | ···················· | ···················· | ···················· |
| H | 4276 | hCoV-19/India/TG-CCMB-AA335/2020\|EPI_ISL_539756\|20 | ···················· | ···················· | ···················· | ···················· | ···················· | ···················· |
| H | 4277 | hCoV-19/India/TG-CCMB-AA372/2020\|EPI_ISL_539757\|20 | ···················· | ···················· | ···················· | ···················· | ···················· | ···················· |
| H | 4278 | hCoV-19/India/TG-CCMB-AA42/2020\|EPI_ISL_539758\|202 | ···················· | ···················· | ···T················ | ···················· | ···················· | ···················· |
| H | 4279 | hCoV-19/India/TG-CCMB-AA617/2020\|EPI_ISL_539759\|20 | ···················· | ···················· | ···················· | ···················· | ···················· | ···················· |
| H | 4280 | hCoV-19/India/TG-CCMB-AA636/2020\|EPI_ISL_539760\|20 | ···················· | ···················· | ···················· | ···················· | ···················· | ···················· |
| H | 4281 | hCoV-19/India/TG-CCMB-AA68/2020\|EPI_ISL_539761\|202 | ···················· | ···················· | ···················· | ···················· | ···················· | ···················· |
| H | 4282 | hCoV-19/India/TG-CCMB-AA833/2020\|EPI_ISL_539762\|20 | ···················· | ···················· | ···················· | ···················· | ···················· | ···················· |
| H | 4283 | hCoV-19/India/TG-CCMB-AA979/2020\|EPI_ISL_539763\|20 | ···················· | ···················· | ···················· | ···················· | ···················· | ···················· |
| H | 4284 | hCoV-19/India/TG-CCMB-AB229/2020\|EPI_ISL_539764\|20 | ···················· | ···················· | ···················· | ···················· | ···················· | ···················· |
| H | 4285 | hCoV-19/India/TG-CCMB-AB247/2020\|EPI_ISL_539765\|20 | ···················· | ···················· | ···················· | ···················· | ···················· | ···················· |
| H | 4286 | hCoV-19/India/TG-CCMB-AB249/2020\|EPI_ISL_539766\|20 | ···················· | ···················· | ···················· | ···················· | ···················· | ···················· |
| H | 4287 | hCoV-19/India/TG-CCMB-AB297/2020\|EPI_ISL_539767\|20 | ···················· | ···················· | ···················· | ···················· | ···················· | ···················· |
| H | 4288 | hCoV-19/India/TG-CCMB-AB430/2020\|EPI_ISL_539768\|20 | ···················· | ···················· | ···················· | ···················· | ···················· | ···················· |
| H | 4289 | hCoV-19/India/TG-CCMB-AB490/2020\|EPI_ISL_539769\|20 | ···················· | ···················· | ···················· | ···················· | ···················· | ···················· |
| H | 4290 | hCoV-19/India/TG-CCMB-AB519/2020\|EPI_ISL_539770\|20 | ···················· | ···················· | ···················· | ···················· | ···················· | ···················· |
| H | 4291 | hCoV-19/India/TG-CCMB-AB523/2020\|EPI_ISL_539771\|20 | ···················· | ···················· | ···················· | ···················· | ···················· | ···················· |
| H | 4292 | hCoV-19/India/TG-CCMB-AC529/2020\|EPI_ISL_539772\|20 | ···················· | ···················· | ···················· | ···················· | ···················· | ···················· |
| H | 4293 | hCoV-19/India/TG-CCMB-AC576/2020\|EPI_ISL_539773\|20 | ···················· | ···················· | ···················· | ···················· | ···················· | ···················· |
| H | 4294 | hCoV-19/India/TG-CCMB-R124/2020\|EPI_ISL_539774\|202 | ···················· | ···················· | ···················· | ···················· | ···················· | ···················· |
| H | 4295 | hCoV-19/India/TG-CCMB-R325/2020\|EPI_ISL_539775\|202 | ···················· | ···················· | ···················· | ···················· | ···················· | ···················· |
| H | 4296 | hCoV-19/Czech Republic/NRL-5607/2020\|EPI_ISL_53977 | ···················· | ···················· | ···················· | ···················· | ···················· | ···················· |
| H | 4297 | hCoV-19/Czech Republic/NRL-5751/2020\|EPI_ISL_53977 | ···················· | ···················· | ···················· | ···················· | ···················· | ···················· |
| H | 4298 | hCoV-19/Czech Republic/NRL-5886-1/2020\|EPI_ISL_539 | ···················· | ···················· | ···················· | ···················· | ···················· | ···················· |
| H | 4299 | hCoV-19/Czech Republic/NRL-5886-2/2020\|EPI_ISL_539 | ···················· | ···················· | ···················· | ···················· | ···················· | ···················· |
| H | 4300 | hCoV-19/Czech Republic/NRL-6047/2020\|EPI_ISL_53978 | ···················· | ···················· | ···················· | ···················· | ···················· | ···················· |
| H | 4301 | hCoV-19/Czech Republic/NRL-3968/2020\|EPI_ISL_53978 | ···················· | ···················· | ···················· | ···················· | ···················· | ···················· |
| H | 4302 | hCoV-19/Czech Republic/NRL-6832/2020\|EPI_ISL_53978 | ···················· | ···················· | ···················· | ···················· | ···················· | ···················· |
| H | 4303 | hCoV-19/Ecuador/USFQ-208/2020\|EPI_ISL_539783\|2020- | ···················· | ···················· | ···················· | ···················· | ···················· | ···················· |
| H | 4304 | hCoV-19/Ecuador/USFQ-209/2020\|EPI_ISL_539784\|2020- | ···················· | ···················· | ···················· | ···················· | ···················· | ···················· |
| H | 4305 | hCoV-19/Ecuador/USFQ-212/2020\|EPI_ISL_539785\|2020- | ···················· | ···················· | ···················· | ···················· | ···················· | ···················· |
| H | 4306 | hCoV-19/Ecuador/USFQ-213/2020\|EPI_ISL_539786\|2020- | ···················· | ···················· | ···················· | ···················· | ···················· | ···················· |
| H | 4307 | hCoV-19/Ecuador/USFQ-214/2020\|EPI_ISL_539787\|2020- | ···················· | ···················· | ···················· | ···················· | ···················· | ···················· |
| H | 4308 | hCoV-19/Ecuador/USFQ-216/2020\|EPI_ISL_539788\|2020- | ···················· | ···················· | ···················· | ···················· | ···················· | ···················· |
| H | 4309 | hCoV-19/Ecuador/USFQ-219/2020\|EPI_ISL_539789\|2020- | ···················· | ···················· | ···················· | ···················· | ···················· | ···················· |
| H | 4310 | hCoV-19/Ecuador/USFQ-220/2020\|EPI_ISL_539790\|2020- | ···················· | ···················· | ···················· | ···················· | ···················· | ···················· |
| H | 4311 | hCoV-19/Ecuador/USFQ-222/2020\|EPI_ISL_539791\|2020- | ···················· | ···················· | ···················· | ···················· | ···················· | ···················· |
| H | 4312 | hCoV-19/Ecuador/USFQ-224/2020\|EPI_ISL_539792\|2020- | ···················· | ···················· | ···················· | ···················· | ···················· | ···················· |
| H | 4313 | hCoV-19/Ecuador/USFQ-226/2020\|EPI_ISL_539793\|2020- | ···················· | ···················· | ···················· | ···················· | ···················· | ···················· |
| H | 4314 | hCoV-19/USA/WY-WYPHL-20008408/2020\|EPI_ISL_539794\| | ···················· | ···················· | ···················· | ···················· | ···················· | ···················· |
| H | 4315 | hCoV-19/USA/WY-WYPHL-20008859/2020\|EPI_ISL_539795\| | ···················· | ···················· | ···················· | ···················· | ···················· | ···················· |
| H | 4316 | hCoV-19/USA/WY-WYPHL-20009305/2020\|EPI_ISL_539796\| | ···················· | ···················· | ···················· | ···················· | ···················· | ···················· |
| H | 4317 | hCoV-19/USA/WY-WYPHL-20009722/2020\|EPI_ISL_539797\| | ···················· | ···················· | ···················· | ···················· | ···················· | ···················· |
| H | 4318 | hCoV-19/USA/WY-WYPHL-20009973/2020\|EPI_ISL_539798\| | ···················· | ···················· | ···················· | ···················· | ···················· | ···················· |
| H | 4319 | hCoV-19/USA/WY-WYPHL-20010244/2020\|EPI_ISL_539799\| | ···················· | ···················· | ···················· | ···················· | ···················· | ···················· |
| H | 4320 | hCoV-19/England/SHEF-C184F/2020\|EPI_ISL_540720\|202 | ···················· | ···················· | ···················· | ···················· | ···················· | ···················· |
| H | 4321 | hCoV-19/England/SHEF-C1988/2020\|EPI_ISL_540721\|202 | ···················· | ···················· | ···················· | ···················· | ···················· | ···················· |
| H | 4322 | hCoV-19/England/SHEF-C24FF/2020\|EPI_ISL_540722\|202 | ···················· | ···················· | ···················· | ···················· | ···················· | ···················· |
| H | 4323 | hCoV-19/England/SHEF-C483D/2020\|EPI_ISL_540728\|202 | ···················· | ···················· | ···················· | ···················· | ···················· | ···················· |
| H | 4324 | hCoV-19/England/SHEF-C509E/2020\|EPI_ISL_540729\|202 | ···················· | ···················· | ···················· | ···················· | ···················· | ···················· |
| H | 4325 | hCoV-19/England/SHEF-C5896/2020\|EPI_ISL_540732\|202 | ···················· | ···················· | ···················· | ···················· | ···················· | ···················· |
| H | 4326 | hCoV-19/England/SHEF-C5D5B/2020\|EPI_ISL_540733\|202 | ···················· | ···················· | ···················· | ···················· | ···················· | ···················· |
| H | 4327 | hCoV-19/England/SHEF-C6CD5/2020\|EPI_ISL_540734\|202 | ···················· | ···················· | ···················· | ···················· | ···················· | ···················· |
| H | 4328 | hCoV-19/England/SHEF-C6E75/2020\|EPI_ISL_540735\|202 | ···················· | ···················· | ···················· | ···················· | ···················· | ···················· |
| H | 4329 | hCoV-19/England/SHEF-C7111/2020\|EPI_ISL_540736\|202 | ···················· | ···················· | ···················· | ···················· | ···················· | ···················· |
| H | 4330 | hCoV-19/England/SHEF-C955F/2020\|EPI_ISL_540743\|202 | ···················· | ···················· | ···················· | ···················· | ···················· | ···················· |
| H | 4331 | hCoV-19/England/SHEF-C96D4/2020\|EPI_ISL_540744\|202 | ···················· | ···················· | ···················· | ···················· | ···················· | ···················· |
| H | 4332 | hCoV-19/England/SHEF-C9D2A/2020\|EPI_ISL_540746\|202 | ···················· | ···················· | ···················· | ···················· | ···················· | ···················· |
| H | 4333 | hCoV-19/England/SHEF-CAA31/2020\|EPI_ISL_540751\|202 | ···················· | ···················· | ···················· | ···················· | ···················· | ···················· |
| H | 4334 | hCoV-19/England/SHEF-CAE62/2020\|EPI_ISL_540752\|202 | ···················· | ···················· | ···················· | ···················· | ···················· | ···················· |
| H | 4335 | hCoV-19/England/SHEF-CB03E/2020\|EPI_ISL_540753\|202 | ···················· | ···················· | ···················· | ···················· | ···················· | ···················· |
| H | 4336 | hCoV-19/England/SHEF-CDDAE/2020\|EPI_ISL_540756\|202 | ···················· | ···················· | ···················· | ···················· | ···················· | ···················· |
| H | 4337 | hCoV-19/England/SHEF-CE8BB/2020\|EPI_ISL_540758\|202 | ···················· | ···················· | ···················· | ···················· | ···················· | ···················· |
| H | 4338 | hCoV-19/England/SHEF-CF243/2020\|EPI_ISL_540760\|202 | ···················· | ···················· | ···················· | ···················· | ···················· | ···················· |
| H | 4339 | hCoV-19/England/SHEF-D0C6F/2020\|EPI_ISL_540767\|202 | ···················· | ···················· | ···················· | ···················· | ···················· | ···················· |
| H | 4340 | hCoV-19/England/SHEF-D1141/2020\|EPI_ISL_540768\|202 | ···················· | ···················· | ···················· | ···················· | ···················· | ···················· |
| H | 4341 | hCoV-19/England/SHEF-D1F38/2020\|EPI_ISL_540770\|202 | ···················· | ···················· | ···················· | ···················· | ···················· | ···················· |
| H | 4342 | hCoV-19/Scotland/CVR3978/2020\|EPI_ISL_540775\|2020- | ···················· | ···················· | ···················· | ···················· | ···················· | ···················· |

| | | | | | | | | |
|---|---|---|---|---|---|---|---|---|
| H | 4343 hCoV-19/Scotland/CVR3980/2020\|EPI_ISL_540777\|2020- | ···················· | ···················· | ···················· | ···················· | ···················· | ···················· | |
| H | 4344 hCoV-19/Scotland/CVR3981/2020\|EPI_ISL_540778\|2020- | ···················· | ···················· | ···················· | ···················· | ···················· | ···················· | |
| H | 4345 hCoV-19/Scotland/CVR3982/2020\|EPI_ISL_540779\|2020- | ···················· | ···················· | ···················· | ···················· | ···················· | ···················· | |
| H | 4346 hCoV-19/Scotland/CVR3983/2020\|EPI_ISL_540780\|2020- | ···················· | ···················· | ···················· | ···················· | ···················· | ···················· | |
| H | 4347 hCoV-19/Scotland/CVR3987/2020\|EPI_ISL_540784\|2020- | ···················· | ···················· | ···················· | ···················· | ···················· | ···················· | |
| H | 4348 hCoV-19/Scotland/CVR3988/2020\|EPI_ISL_540785\|2020- | ···················· | ···················· | ···················· | ···················· | ···················· | ···················· | |
| H | 4349 hCoV-19/Scotland/CVR3989/2020\|EPI_ISL_540786\|2020- | ···················· | ···················· | ···················· | ···················· | ···················· | ···················· | |
| H | 4350 hCoV-19/Scotland/CVR3990/2020\|EPI_ISL_540787\|2020- | ···················· | ···················· | ···················· | ···················· | ···················· | ···················· | |
| H | 4351 hCoV-19/Scotland/CVR3992/2020\|EPI_ISL_540789\|2020- | ···················· | ···················· | ···················· | ···················· | ···················· | ···················· | |
| H | 4352 hCoV-19/Scotland/CVR3994/2020\|EPI_ISL_540791\|2020- | ···················· | ···················· | ···················· | ···················· | ···················· | ···················· | |
| H | 4353 hCoV-19/England/QEUH-99046A/2020\|EPI_ISL_530374\|20 | ···················· | ···················· | ···················· | ···················· | ···················· | ···················· | |
| H | 4354 hCoV-19/England/QEUH-99027F/2020\|EPI_ISL_530375\|20 | ···················· | ···················· | ···················· | ···················· | ···················· | ···················· | |
| H | 4355 hCoV-19/England/QEUH-990734/2020\|EPI_ISL_530378\|20 | ···················· | ···················· | ···················· | ···················· | ···················· | ···················· | |
| H | 4356 hCoV-19/England/QEUH-98FD63/2020\|EPI_ISL_530380\|20 | ···················· | ·A·················· | ···················· | ···················· | ···················· | ···················· | |
| H | 4357 hCoV-19/England/QEUH-990743/2020\|EPI_ISL_530382\|20 | ···················· | ···················· | ···················· | ···················· | ···················· | ···················· | |
| H | 4358 hCoV-19/England/QEUH-98FC75/2020\|EPI_ISL_530383\|20 | ···················· | ···················· | ···················· | ···················· | ···················· | ···················· | |
| H | 4359 hCoV-19/England/QEUH-9904F1/2020\|EPI_ISL_530384\|20 | ···················· | ·A·················· | ···················· | ···················· | ···················· | ···················· | |
| H | 4360 hCoV-19/Scotland/QEUH-9902D9/2020\|EPI_ISL_530386\|2 | ···················· | ···················· | ···················· | ···················· | ···················· | ···················· | |
| H | 4361 hCoV-19/England/QEUH-98FBA5/2020\|EPI_ISL_530387\|20 | ···················· | ···················· | ···················· | ···················· | ···················· | ···················· | |
| H | 4362 hCoV-19/England/QEUH-990716/2020\|EPI_ISL_530390\|20 | ···················· | ···················· | ···················· | ···················· | ···················· | ···················· | |
| H | 4363 hCoV-19/Northern Ireland/QEUH-96EC8B/2020\|EPI_ISL_ | ···················· | ···················· | ···················· | ···················· | ···················· | ···················· | |
| H | 4364 hCoV-19/England/QEUH-96E104/2020\|EPI_ISL_530395\|20 | ···················· | ···················· | ···················· | ···················· | ···················· | ···················· | |
| H | 4365 hCoV-19/England/QEUH-96F17C/2020\|EPI_ISL_530396\|20 | ···················· | ···················· | ···················· | ···················· | ···················· | ···················· | |
| H | 4366 hCoV-19/Northern Ireland/QEUH-96EACD/2020\|EPI_ISL_ | ···················· | ···················· | ···················· | ···················· | ···················· | ···················· | |
| H | 4367 hCoV-19/England/QEUH-96E4A1/2020\|EPI_ISL_530399\|20 | ···················· | ···················· | ···················· | ···················· | ···················· | ···················· | |
| H | 4368 hCoV-19/England/QEUH-96E6D8/2020\|EPI_ISL_530400\|20 | ···················· | ···················· | ···················· | ···················· | ···················· | ···················· | |
| H | 4369 hCoV-19/England/QEUH-963688/2020\|EPI_ISL_531698\|20 | ···················· | ···················· | ···················· | ···················· | ···················· | ···················· | |
| H | 4370 hCoV-19/England/QEUH-96270E/2020\|EPI_ISL_531699\|20 | ···················· | ···················· | ···················· | ···················· | ···················· | ···················· | |
| H | 4371 hCoV-19/England/QEUH-962BF3/2020\|EPI_ISL_531731\|20 | ···················· | ···················· | ···················· | ···················· | ···················· | ···················· | |
| H | 4372 hCoV-19/Scotland/QEUH-961F61/2020\|EPI_ISL_531732\|2 | ···················· | ···················· | ···················· | ···················· | ···················· | ···················· | |
| H | 4373 hCoV-19/Wales/PHWC-16A0F4/2020\|EPI_ISL_535201\|2020 | ···················· | ·A·················· | ···················· | ···················· | ···················· | ··A················· | |
| H | 4374 hCoV-19/Wales/PHWC-16A11F/2020\|EPI_ISL_535203\|2020 | ···················· | ···················· | ···················· | ···················· | ···················· | ···················· | |
| H | 4375 hCoV-19/Wales/PHWC-16A12E/2020\|EPI_ISL_535204\|2020 | ···················· | ···················· | ···················· | ···················· | ···················· | ···················· | |
| H | 4376 hCoV-19/Wales/PHWC-16A14C/2020\|EPI_ISL_535206\|2020 | ···················· | ···················· | ···················· | ···················· | ···················· | ···················· | |
| H | 4377 hCoV-19/Wales/PHWC-16A16A/2020\|EPI_ISL_535207\|2020 | ···················· | ···················· | ···················· | ···················· | ···················· | ···················· | |
| H | 4378 hCoV-19/Wales/PHWC-16A179/2020\|EPI_ISL_535208\|2020 | ···················· | ···················· | ···················· | ···················· | ···················· | ···················· | |
| H | 4379 hCoV-19/Wales/PHWC-16A188/2020\|EPI_ISL_535209\|2020 | ···················· | ···················· | ···················· | ···················· | ···················· | ···················· | |
| H | 4380 hCoV-19/Wales/PHWC-16A197/2020\|EPI_ISL_535210\|2020 | ···················· | ···················· | ···················· | ···················· | ···················· | ···················· | |
| H | 4381 hCoV-19/Wales/PHWC-16A1B5/2020\|EPI_ISL_535212\|2020 | ···················· | ···················· | ···················· | ···················· | ···················· | ···················· | |
| H | 4382 hCoV-19/Wales/PHWC-16A1C4/2020\|EPI_ISL_535213\|2020 | ···················· | ···················· | ···················· | ···················· | ···················· | ···················· | |
| H | 4383 hCoV-19/Wales/PHWC-16A1E2/2020\|EPI_ISL_535214\|2020 | ···················· | ···················· | ···················· | ···················· | ···················· | ···················· | |
| H | 4384 hCoV-19/Wales/PHWC-16A1F1/2020\|EPI_ISL_535215\|2020 | ···················· | ···················· | ···················· | ···················· | ···················· | ···················· | |
| H | 4386 hCoV-19/Wales/PHWC-16A22B/2020\|EPI_ISL_535218\|2020 | ···················· | ···················· | ···················· | ···················· | ···················· | ···················· | |
| H | 4387 hCoV-19/Wales/PHWC-16A23A/2020\|EPI_ISL_535219\|2020 | ···················· | ···················· | ···················· | ···················· | ···················· | ···················· | |
| H | 4388 hCoV-19/Wales/PHWC-16A249/2020\|EPI_ISL_535220\|2020 | ···················· | ···················· | ···················· | ···················· | ···················· | ···················· | |
| H | 4390 hCoV-19/Wales/PHWC-16A2A3/2020\|EPI_ISL_535226\|2020 | ···················· | ···················· | ···················· | ···················· | ···················· | ···················· | |
| H | 4392 hCoV-19/Wales/PHWC-16A2C1/2020\|EPI_ISL_535228\|2020 | ···················· | ···················· | ···················· | ···················· | ···················· | ···················· | |
| H | 4394 hCoV-19/Wales/PHWC-16A319/2020\|EPI_ISL_535233\|2020 | ···················· | ···················· | ···················· | ···················· | ···················· | ···················· | |
| H | 4396 hCoV-19/Wales/PHWC-16A346/2020\|EPI_ISL_535236\|2020 | ···················· | ···················· | ···················· | ···················· | ···················· | ···················· | |
| H | 4398 hCoV-19/Wales/PHWC-16A373/2020\|EPI_ISL_535239\|2020 | ···················· | ···················· | ···················· | ···················· | ···················· | ···················· | |
| H | 4400 hCoV-19/Wales/PHWC-16A3EC/2020\|EPI_ISL_535245\|2020 | ···················· | ···················· | ···················· | ···················· | ···················· | ···················· | |
| H | 4404 hCoV-19/Wales/PHWC-16A425/2020\|EPI_ISL_535249\|2020 | ···················· | ···················· | ···················· | ···················· | NNN················· | ···················· | 1 |
| H | 4405 hCoV-19/Wales/PHWC-16A434/2020\|EPI_ISL_535250\|2020 | ···················· | ···················· | ···················· | ···················· | ···················· | ···················· | |
| H | 4406 hCoV-19/Wales/PHWC-16A443/2020\|EPI_ISL_535251\|2020 | ···················· | ···················· | ···················· | ···················· | ···················· | ···················· | |
| H | 4407 hCoV-19/Wales/PHWC-16A470/2020\|EPI_ISL_535254\|2020 | ···················· | ···················· | ···················· | ···················· | ···················· | ···················· | |
| H | 4408 hCoV-19/Wales/PHWC-16A4AD/2020\|EPI_ISL_535257\|2020 | ···················· | ···················· | ···················· | ···················· | ···················· | ···················· | |
| H | 4409 hCoV-19/Peru/LIM-INS-101/2020\|EPI_ISL_536500\|2020- | ···················· | ···················· | ···················· | ···················· | ···················· | ···················· | |
| H | 4410 hCoV-19/Peru/CAL-INS-103/2020\|EPI_ISL_536502\|2020- | ···················· | ···················· | ···················· | ···················· | ···················· | ···················· | |
| H | 4411 hCoV-19/Peru/CAL-INS-104/2020\|EPI_ISL_536503\|2020- | ···················· | ···················· | ···················· | ···················· | ···················· | ···················· | |
| H | 4412 hCoV-19/Peru/CAL-INS-105/2020\|EPI_ISL_536504\|2020- | ···················· | ···················· | ···················· | ···················· | ···················· | ···················· | |
| H | 4413 hCoV-19/Peru/ICA-INS-107/2020\|EPI_ISL_536505\|2020- | ···················· | ···················· | ···················· | ···················· | ···················· | ···················· | |
| H | 4414 hCoV-19/Peru/ICA-INS-108/2020\|EPI_ISL_536506\|2020- | ···················· | ···················· | ···················· | ···················· | ···················· | ···················· | |
| H | 4415 hCoV-19/Peru/LIM-INS-110/2020\|EPI_ISL_536508\|2020- | ···················· | ···················· | ···················· | ···················· | ···················· | ···················· | |
| H | 4416 hCoV-19/Peru/LIM-INS-111/2020\|EPI_ISL_536509\|2020- | ···················· | ···················· | ···················· | ···················· | ···················· | ···················· | |
| H | 4417 hCoV-19/Peru/LIM-INS-112/2020\|EPI_ISL_536510\|2020- | ···················· | ···················· | ···················· | ···················· | ···················· | ···················· | |
| H | 4418 hCoV-19/Peru/LIM-INS-113/2020\|EPI_ISL_536511\|2020- | ···················· | ···················· | ···················· | ···················· | ···················· | ···················· | |
| H | 4419 hCoV-19/Peru/LIM-INS-114/2020\|EPI_ISL_536512\|2020- | ···················· | ···················· | ···················· | ···················· | ···················· | ···················· | |
| H | 4420 hCoV-19/Peru/LIM-INS-115/2020\|EPI_ISL_536513\|2020- | ···················· | ···················· | ···················· | ···················· | ···················· | ···················· | |
| H | 4421 hCoV-19/Peru/LIM-INS-116/2020\|EPI_ISL_536514\|2020- | ···················· | ···················· | ···················· | ···················· | ···················· | ···················· | |
| H | 4422 hCoV-19/Peru/LIM-INS-117/2020\|EPI_ISL_536515\|2020- | ···················· | ···················· | ···················· | ···················· | ···················· | ···················· | |
| H | 4423 hCoV-19/Peru/LIM-INS-118/2020\|EPI_ISL_536516\|2020- | ···················· | ···················· | ···················· | ···················· | ···················· | ···················N | 1 |
| H | 4424 hCoV-19/Peru/LIM-INS-119/2020\|EPI_ISL_536517\|2020- | ···················· | ···················· | ···················· | ···················· | ···················· | ···················· | |
| H | 4425 hCoV-19/Peru/LIM-INS-120/2020\|EPI_ISL_536518\|2020- | ···················· | ···················· | ···················· | ···················· | ···················· | ···················· | |
| H | 4426 hCoV-19/Peru/LIM-INS-121/2020\|EPI_ISL_536519\|2020- | ···················· | ···················· | ···················· | ···················· | ···················· | ···················· | |
| H | 4427 hCoV-19/Peru/LIM-INS-122/2020\|EPI_ISL_536520\|2020- | ···················· | ···················· | ···················· | ···················· | ···················· | ···················· | |
| H | 4428 hCoV-19/Peru/LIM-INS-123/2020\|EPI_ISL_536521\|2020- | ···················· | ···················· | ···················· | ···················· | ···················· | ···················· | |
| H | 4429 hCoV-19/Peru/LIM-INS-124/2020\|EPI_ISL_536522\|2020- | ···················· | ···················· | ···················· | ···················· | ···················· | ···················· | |
| H | 4430 hCoV-19/Peru/LAM-INS-125/2020\|EPI_ISL_536523\|2020- | ···················· | ···················· | ···················· | ···················· | ···················· | ···················· | |
| H | 4431 hCoV-19/Peru/LAM-INS-126/2020\|EPI_ISL_536524\|2020- | ···················· | ···················· | ···················· | ···················· | ···················· | ···················· | |
| H | 4432 hCoV-19/Peru/LIM-INS-130/2020\|EPI_ISL_536527\|2020- | ···················· | ···················· | ···················· | ···················· | ···················· | ···················· | |

| | | | | | | | | | |
|---|---|---|---|---|---|---|---|---|---|
| H | 4433 | hCoV-19/Peru/JUN-INS-131/2020\|EPI_ISL_536528\|2020- | .................... | .................... | ..................... | .................... | .................... | .................... | |
| H | 4434 | hCoV-19/Peru/ANC-INS-132/2020\|EPI_ISL_536529\|2020- | .................... | .................... | ..................... | .................... | .................... | .................... | |
| H | 4435 | hCoV-19/Peru/LIM-INS-136/2020\|EPI_ISL_536531\|2020- | .................... | .................... | ..................... | .................... | .................... | .................... | |
| H | 4436 | hCoV-19/Peru/LIM-INS-138/2020\|EPI_ISL_536533\|2020- | .................... | .................... | ..................... | .................... | .................... | .................... | |
| H | 4437 | hCoV-19/Peru/LIM-INS-139/2020\|EPI_ISL_536534\|2020- | .................... | .................... | ..................... | .................... | .................... | .................... | |
| H | 4438 | hCoV-19/Peru/LIM-INS-142/2020\|EPI_ISL_536537\|2020- | .................... | .................... | ..................... | .................... | .................... | .................... | |
| H | 4439 | hCoV-19/Peru/LIM-INS-143/2020\|EPI_ISL_536538\|2020- | .................... | .................... | ..................... | .................... | .................... | .................... | |
| H | 4440 | hCoV-19/Peru/LIM-INS-144/2020\|EPI_ISL_536539\|2020- | .................... | .................... | ..................... | .................... | .................... | .................... | |
| H | 4441 | hCoV-19/Peru/LIM-INS-145/2020\|EPI_ISL_536540\|2020- | .................... | .................... | ..................... | .................... | .................... | .................... | |
| H | 4442 | hCoV-19/Peru/LIM-INS-146/2020\|EPI_ISL_536541\|2020- | .................... | .................... | ..................... | .................... | .................... | .................... | |
| H | 4443 | hCoV-19/Peru/LIM-INS-147/2020\|EPI_ISL_536542\|2020- | .................... | .................... | ..................... | .................... | .................... | .................... | |
| H | 4444 | hCoV-19/Peru/LIM-INS-148/2020\|EPI_ISL_536543\|2020- | .................... | .................... | ..................... | .................... | .................... | .................... | |
| H | 4445 | hCoV-19/Peru/LIM-INS-150/2020\|EPI_ISL_536545\|2020- | .................... | .................... | ..................... | .................... | .................... | .................... | |
| H | 4446 | hCoV-19/Peru/LIM-INS-155/2020\|EPI_ISL_536548\|2020- | .................... | .................... | ..................... | .................... | .................... | .................... | |
| H | 4447 | hCoV-19/Peru/LIM-INS-156/2020\|EPI_ISL_536549\|2020- | .................... | .................... | ..................... | .................... | .................... | .................... | |
| H | 4448 | hCoV-19/Peru/LIM-INS-157/2020\|EPI_ISL_536550\|2020- | .................... | .................... | ..................... | .................... | .................... | .................... | |
| H | 4449 | hCoV-19/Peru/LIM-INS-158/2020\|EPI_ISL_536551\|2020- | .................... | .................... | ..................... | .................... | .................... | .................... | |
| H | 4450 | hCoV-19/Peru/LIM-INS-159/2020\|EPI_ISL_536552\|2020- | .................... | .................... | ..................... | .................... | .................... | .................... | |
| H | 4451 | hCoV-19/Peru/LIM-INS-160/2020\|EPI_ISL_536553\|2020- | .................... | .................... | ..................... | .................... | .................... | .................... | |
| H | 4452 | hCoV-19/Peru/LIM-INS-161/2020\|EPI_ISL_536554\|2020- | .................... | .................... | ..................... | .................... | .................... | .................... | |
| H | 4453 | hCoV-19/Peru/LIM-INS-162/2020\|EPI_ISL_536555\|2020- | .................... | .................... | ..................... | .................... | .................... | .................... | |
| H | 4454 | hCoV-19/Peru/CAL-INS-163/2020\|EPI_ISL_536556\|2020- | .................... | .................... | ..................... | .................... | .................... | .................... | |
| H | 4455 | hCoV-19/Peru/ARE-INS-164/2020\|EPI_ISL_536557\|2020- | .................... | .................... | ..................... | .................... | .................... | .................... | |
| H | 4456 | hCoV-19/Peru/ARE-INS-165/2020\|EPI_ISL_536558\|2020- | .................... | .................... | ..................... | .................... | .................... | .................... | |
| H | 4457 | hCoV-19/Peru/ICA-INS-166/2020\|EPI_ISL_536559\|2020- | .................... | .................... | ..................... | .................... | .................... | .................... | |
| H | 4458 | hCoV-19/Peru/LIM-INS-167/2020\|EPI_ISL_536560\|2020- | .................... | .................... | ..................... | .................... | .................... | .................... | |
| H | 4459 | hCoV-19/Peru/LIM-INS-168/2020\|EPI_ISL_536561\|2020- | .................... | .................... | ..................... | .................... | .................... | .................... | |
| H | 4460 | hCoV-19/Peru/LAM-INS-169/2020\|EPI_ISL_536562\|2020- | .................... | .................... | ..................... | .................... | .................... | .................... | |
| H | 4461 | hCoV-19/USA/WI-UW-1199/2020\|EPI_ISL_536572\|2020-08 | .................... | .................... | ..................... | .................... | .................... | .................... | |
| H | 4462 | hCoV-19/USA/WI-UW-1200/2020\|EPI_ISL_536573\|2020-08 | .................... | .................... | ..................... | .................... | .................... | .................... | |
| H | 4463 | hCoV-19/USA/WI-UW-1201/2020\|EPI_ISL_536574\|2020-07 | .................... | .................... | ..................... | .................... | .................... | .................... | |
| H | 4464 | hCoV-19/USA/WI-UW-1202/2020\|EPI_ISL_536575\|2020-08 | .................... | .................... | ..................... | .................... | .................... | .................... | |
| H | 4465 | hCoV-19/USA/WI-UW-1203/2020\|EPI_ISL_536576\|2020-08 | .................... | .................... | ..................... | .................... | .................... | .................... | |
| H | 4466 | hCoV-19/USA/WI-UW-1204/2020\|EPI_ISL_536577\|2020-08 | .................... | ..................C. | ..................... | .................... | .................... | .................... | 1 |
| H | 4467 | hCoV-19/USA/WI-UW-1205/2020\|EPI_ISL_536578\|2020-07 | .................... | .................... | ..................... | .................... | .................... | .................... | |
| H | 4468 | hCoV-19/USA/WI-UW-1206/2020\|EPI_ISL_536579\|2020-08 | .................... | .................... | ..................... | .................... | .................... | .................... | |
| H | 4469 | hCoV-19/USA/WI-UW-1207/2020\|EPI_ISL_536580\|2020-08 | .................... | .................... | ..................... | .................... | .................... | .................... | |
| H | 4470 | hCoV-19/USA/WI-UW-1208/2020\|EPI_ISL_536581\|2020-08 | .................... | .................... | ..................... | .................... | .................... | .................... | |
| H | 4471 | hCoV-19/USA/WI-UW-1209/2020\|EPI_ISL_536582\|2020-08 | .................... | .................... | ..................... | .................... | .................... | .................... | |
| H | 4472 | hCoV-19/USA/WI-UW-1210/2020\|EPI_ISL_536583\|2020-08 | .................... | .................... | ..................... | .................... | .................... | .................... | |
| H | 4473 | hCoV-19/USA/WI-UW-1211/2020\|EPI_ISL_536584\|2020-08 | .................... | .................... | ..................... | .................... | .................... | .................... | |
| H | 4474 | hCoV-19/USA/WI-UW-1212/2020\|EPI_ISL_536585\|2020-08 | .................... | .................... | ..................... | .................... | .................... | .................... | |
| H | 4475 | hCoV-19/USA/WI-UW-1213/2020\|EPI_ISL_536586\|2020-08 | .................... | .................... | ..................... | .................... | .................... | .................... | |
| H | 4476 | hCoV-19/USA/WI-UW-1214/2020\|EPI_ISL_536587\|2020-08 | .................... | .................... | ..................... | .................... | .................... | .................... | |
| H | 4477 | hCoV-19/USA/WI-UW-1215/2020\|EPI_ISL_536588\|2020-08 | .................... | .............A...... | ..................... | .................... | .................... | .................... | |
| H | 4479 | hCoV-19/England/QEUH-9618FD/2020\|EPI_ISL_537146\|20 | .................... | .................... | ..................... | .................... | .................... | .................... | |
| H | 4480 | hCoV-19/England/QEUH-99824A/2020\|EPI_ISL_537148\|20 | .................... | .................... | ..................... | .................... | .................... | .................... | |
| H | 4481 | hCoV-19/England/QEUH-997FF9/2020\|EPI_ISL_537150\|20 | .................... | .................... | ..................... | .................... | ....T............... | .................... | |
| H | 4482 | hCoV-19/England/QEUH-997D86/2020\|EPI_ISL_537151\|20 | .................... | .................... | ..................... | .................... | .................... | .................... | |
| H | 4483 | hCoV-19/England/QEUH-997DE0/2020\|EPI_ISL_537153\|20 | .................... | A................... | ..................... | .................... | .................... | .................... | |
| H | 4484 | hCoV-19/England/QEUH-997F17/2020\|EPI_ISL_537154\|20 | .................... | .................... | ..................... | .................... | .................... | .................... | |
| H | 4485 | hCoV-19/England/QEUH-997CE3/2020\|EPI_ISL_537156\|20 | .................... | .................... | ..................... | .................... | .................... | .................... | |
| H | 4486 | hCoV-19/England/QEUH-997E92/2020\|EPI_ISL_537157\|20 | .................... | .................... | ..................... | .................... | .................... | .................... | |
| H | 4487 | hCoV-19/England/QEUH-9981A7/2020\|EPI_ISL_537158\|20 | .................... | .................... | ..................... | .................... | .................... | .................... | |
| H | 4488 | hCoV-19/England/QEUH-997D3B/2020\|EPI_ISL_537159\|20 | .................... | .................... | ..................... | .................... | .................... | .................... | |
| H | 4489 | hCoV-19/Spain/CB-IBV-98002760/2020\|EPI_ISL_537809\| | .................... | .................... | ..................... | .................... | .................... | .................... | |
| H | 4490 | hCoV-19/Spain/CB-IBV-98002761/2020\|EPI_ISL_537810\| | .................... | .................... | ..................... | .................... | .................... | .................... | |
| H | 4491 | hCoV-19/Spain/RI-IBV-98002288/2020\|EPI_ISL_537811\| | .................... | .................... | ..................... | .................... | .NN................. | .................... | |
| H | 4492 | hCoV-19/Spain/RI-IBV-98002299/2020\|EPI_ISL_537822\| | .................... | .................... | ..................... | .................... | .................... | .................... | |
| H | 4493 | hCoV-19/Spain/RI-IBV-98002305/2020\|EPI_ISL_537828\| | .................... | .................... | ..................... | .................... | .................... | .................... | |
| H | 4496 | hCoV-19/Spain/RI-IBV-98002316/2020\|EPI_ISL_537836\| | .................... | .................... | ..................... | .................... | .................... | .................... | |
| H | 4497 | hCoV-19/Spain/RI-IBV-98002317/2020\|EPI_ISL_537837\| | .................... | .................... | ..................... | .................... | .................... | .................... | |
| H | 4500 | hCoV-19/Spain/RI-IBV-98002321/2020\|EPI_ISL_537840\| | .................... | .................... | ..................... | .................... | .................... | .................... | |
| H | 4501 | hCoV-19/Spain/RI-IBV-98002322/2020\|EPI_ISL_537841\| | .................... | .................... | ..................... | .................... | .................... | .................... | |
| H | 4502 | hCoV-19/Spain/RI-IBV-98002323/2020\|EPI_ISL_537842\| | .................... | .................... | ..................... | .................... | .................... | .................... | |
| H | 4511 | hCoV-19/Spain/RI-IBV-98002345/2020\|EPI_ISL_537856\| | .................... | .................... | ..................... | .................... | .................... | .................... | |
| H | 4512 | hCoV-19/Spain/RI-IBV-98002350/2020\|EPI_ISL_537857\| | .................... | .................... | ..................... | .................... | .................... | .................... | |
| H | 4514 | hCoV-19/Spain/RI-IBV-98002358/2020\|EPI_ISL_537859\| | .................... | .................... | ..................... | .................... | .................... | .................... | |
| H | 4519 | hCoV-19/Spain/RI-IBV-98002378/2020\|EPI_ISL_537870\| | .................... | .................... | ..................... | .................... | .................... | .................... | |
| H | 4520 | hCoV-19/Spain/RI-IBV-98002381/2020\|EPI_ISL_537872\| | .................... | .................... | ..................... | .................... | .................... | .................... | |
| H | 4522 | hCoV-19/Singapore/943/2020\|EPI_ISL_538477\|2020-04- | .................... | .................... | ..................... | .................... | .................... | .................... | |
| H | 4523 | hCoV-19/Singapore/944/2020\|EPI_ISL_538478\|2020-04- | .................... | .................... | ..................... | .................... | .................... | .................... | |
| H | 4524 | hCoV-19/Singapore/945/2020\|EPI_ISL_538479\|2020-04- | .................... | .................... | ..................... | .................... | .................... | .................... | |
| H | 4525 | hCoV-19/Singapore/946/2020\|EPI_ISL_538480\|2020-04- | .................... | .................... | ..................... | .................... | .................... | .................... | |
| H | 4526 | hCoV-19/Singapore/947/2020\|EPI_ISL_538481\|2020-04- | .................... | .................... | ..................... | .................... | .................... | .................... | |
| H | 4527 | hCoV-19/Singapore/948/2020\|EPI_ISL_538482\|2020-04- | .................... | .................... | ..................... | .................... | .................... | .................... | |
| H | 4528 | hCoV-19/Singapore/949/2020\|EPI_ISL_538483\|2020-04- | .................... | .................... | ..................... | .................... | .................... | .................... | |
| H | 4529 | hCoV-19/Singapore/950/2020\|EPI_ISL_538484\|2020-04- | .................... | .................... | ..................... | .................... | .................... | .................... | |
| H | 4530 | hCoV-19/Singapore/951/2020\|EPI_ISL_538485\|2020-04- | .................... | .................... | ..................... | .................... | .................... | .................... | |
| H | 4531 | hCoV-19/Singapore/952/2020\|EPI_ISL_538486\|2020-04- | .................... | .................... | ..................... | .................... | .................... | .................... | |

| | | | | | | | | |
|---|---|---|---|---|---|---|---|---|
| H | 4532 | hCoV-19/Singapore/953/2020\|EPI_ISL_538487\|2020-04- | ···················· | ···················· | ···················· | ···················· | ···················· | ···················· |
| H | 4533 | hCoV-19/Singapore/954/2020\|EPI_ISL_538488\|2020-04- | ···················· | ···················· | ···················· | ···················· | ···················· | ···················· |
| H | 4534 | hCoV-19/Singapore/955/2020\|EPI_ISL_538489\|2020-04- | ···················· | ···················· | ···················· | ···················· | ···················· | ···················· |
| H | 4535 | hCoV-19/Scotland/CVR3999/2020\|EPI_ISL_540796\|2020- | ···················· | ···················· | ···················· | ···················· | ···················· | ···················· |
| H | 4536 | hCoV-19/Czech Republic/NRL_7000/2020\|EPI_ISL_54108 | ···················· | ···················· | ···················· | ···················· | ···················· | ···················· |
| H | 4537 | hCoV-19/Italy/LOM-UniMI-L427/2020\|EPI_ISL_542346\|2 | ···················· | ···················· | ···················· | ···················· | ···················· | ···················· |
| H | 4538 | hCoV-19/Italy/LOM-UniMI-L428/2020\|EPI_ISL_542347\|2 | ···················· | ···················· | ···················· | ···················· | ···················· | ···················· |
| H | 4539 | hCoV-19/Italy/LOM-UniMI-L429/2020\|EPI_ISL_542348\|2 | ···················· | ···················· | ···················· | ···················· | ···················· | ···················· |
| H | 4540 | hCoV-19/Italy/LOM-UniMI-L430/2020\|EPI_ISL_542349\|2 | ···················· | ···················· | ···················· | ···················· | ···················· | ···················· |
| H | 4541 | hCoV-19/Italy/LOM-UniMI-L431/2020\|EPI_ISL_542350\|2 | ···················· | ···················· | ···················· | ···················· | ···················· | ···················· |
| H | 4542 | hCoV-19/Italy/LOM-UniMI-L432/2020\|EPI_ISL_542351\|2 | ···················· | ···················· | ···················· | ···················· | ···················· | ···················· |
| H | 4543 | hCoV-19/Italy/LOM-UniMI-L433/2020\|EPI_ISL_542352\|2 | ···················· | ···················· | ···················· | ···················· | ···················· | ···················· |
| H | 4544 | hCoV-19/Italy/LOM-UniMI-L434/2020\|EPI_ISL_542353\|2 | ···················· | ···················· | ···················· | ···················· | ···················· | ···················· |
| H | 4545 | hCoV-19/Italy/LOM-UniMI-L435/2020\|EPI_ISL_542354\|2 | ···················· | ···················· | ···················· | ···················· | ···················· | ···················· |
| H | 4546 | hCoV-19/Italy/LOM-UniMI-L436/2020\|EPI_ISL_542355\|2 | ···················· | ···················· | ···················· | ···················· | ···················· | ···················· |
| H | 4547 | hCoV-19/Italy/LOM-UniMI-L437/2020\|EPI_ISL_542356\|2 | ···················· | ···················· | ···················· | ···················· | ···················· | ···················· |
| H | 4548 | hCoV-19/Italy/LOM-UniMI-L438/2020\|EPI_ISL_542357\|2 | ···················· | ···················· | ···················· | ···················· | ···················· | ···················· |
| H | 4549 | hCoV-19/Italy/LOM-UniMI-L439/2020\|EPI_ISL_542358\|2 | ···················· | ···················· | ···················· | ···················· | ···················· | ···················· |
| H | 4550 | hCoV-19/Italy/LOM-UniMI-L440/2020\|EPI_ISL_542359\|2 | ···················· | ···················· | ···················· | ···················· | ···················· | ···················· |
| H | 4551 | hCoV-19/Italy/LOM-UniMI-L441/2020\|EPI_ISL_542360\|2 | ···················· | ···················· | ···················· | ···················· | ···················· | ···················· |
| H | 4552 | hCoV-19/Italy/LOM-UniMI-L442/2020\|EPI_ISL_542361\|2 | ···················· | ···················· | ···················· | ···················· | ···················· | ···················· |
| H | 4553 | hCoV-19/Italy/LOM-UniMI-L443/2020\|EPI_ISL_542362\|2 | ···················· | ···················· | ···················· | ···················· | ···················· | ···················· |
| H | 4554 | hCoV-19/Italy/LOM-UniMI-L444/2020\|EPI_ISL_542363\|2 | ···················· | ···················· | ···················· | ···················· | ···················· | ···················· |
| H | 4555 | hCoV-19/Italy/LOM-UniMI-L445/2020\|EPI_ISL_542364\|2 | ···················· | ···················· | ···················· | ···················· | ···················· | ···················· |
| H | 4556 | hCoV-19/Italy/LOM-UniMI-L447/2020\|EPI_ISL_542365\|2 | ···················· | ···················· | ···················· | ···················· | ···················· | ···················· |
| H | 4557 | hCoV-19/Italy/LOM-UniMI-L448/2020\|EPI_ISL_542366\|2 | ···················· | ···················· | ···················· | ···················· | ···················· | ···················· |
| H | 4558 | hCoV-19/Italy/LOM-UniMI-L449/2020\|EPI_ISL_542367\|2 | ···················· | ···················· | ···················· | ···················· | ···················· | ···················· |
| H | 4559 | hCoV-19/Italy/LOM-UniMI-L450/2020\|EPI_ISL_542368\|2 | ···················· | ···················· | ···················· | ···················· | ···················· | ···················· |
| H | 4560 | hCoV-19/Italy/LOM-UniMI-L451/2020\|EPI_ISL_542369\|2 | ···················· | ···················· | ···················· | ···················· | ···················· | ···················· |
| H | 4561 | hCoV-19/Italy/LOM-UniMI-L452/2020\|EPI_ISL_542370\|2 | ···················· | ···················· | ···················· | ···················· | ···················· | ···················· |
| H | 4562 | hCoV-19/Italy/LOM-UniMI-L453/2020\|EPI_ISL_542371\|2 | ···················· | ···················· | ···················· | ···················· | ···················· | ···················· |
| H | 4563 | hCoV-19/Italy/LOM-UniMI-L454/2020\|EPI_ISL_542372\|2 | ···················· | ···················· | ···················· | ···················· | ···················· | ···················· |
| H | 4564 | hCoV-19/Italy/LOM-UniMI-L455/2020\|EPI_ISL_542373\|2 | ···················· | ···················· | ···················· | ···················· | ···················· | ···················· |
| H | 4565 | hCoV-19/Italy/LOM-UniMI-L457/2020\|EPI_ISL_542374\|2 | ···················· | ···················· | ···················· | ···················· | ···················· | ···················· |
| H | 4566 | hCoV-19/Italy/LOM-UniMI-L458/2020\|EPI_ISL_542375\|2 | ···················· | ···················· | ···················· | ···················· | ···················· | ···················· |
| H | 4567 | hCoV-19/Italy/LOM-UniMI-L459/2020\|EPI_ISL_542376\|2 | ···················· | ···················· | ···················· | ···················· | ···················· | ···················· |
| H | 4568 | hCoV-19/Italy/LOM-UniMI-L461/2020\|EPI_ISL_542377\|2 | ···················· | ···················· | ···················· | ···················· | ···················· | ···················· |
| H | 4569 | hCoV-19/Italy/LOM-UniMI-L462/2020\|EPI_ISL_542378\|2 | ···················· | ···················· | ···················· | ···················· | ···················· | ···················· |
| H | 4570 | hCoV-19/Italy/LOM-UniMI-L463/2020\|EPI_ISL_542379\|2 | ···················· | ···················· | ···················· | ···················· | ···················· | ···················· |
| H | 4571 | hCoV-19/Italy/LOM-UniMI-L464/2020\|EPI_ISL_542380\|2 | ···················· | ···················· | ···················· | ···················· | ···················· | ···················· |
| H | 4572 | hCoV-19/Italy/LOM-UniMI-L465/2020\|EPI_ISL_542381\|2 | ···················· | ···················· | ···················· | ···················· | ···················· | ···················· |
| H | 4573 | hCoV-19/Italy/LOM-UniMI-L469/2020\|EPI_ISL_542382\|2 | ···················· | ···················· | ···················· | ···················· | ···················· | ···················· |
| H | 4574 | hCoV-19/Italy/LOM-UniMI-L470/2020\|EPI_ISL_542383\|2 | ···················· | ···················· | ···················· | ···················· | ···················· | ···················· |
| H | 4575 | hCoV-19/Italy/LOM-UniMI-L471/2020\|EPI_ISL_542384\|2 | ···················· | ···················· | ···················· | ···················· | ···················· | ···················· |
| H | 4576 | hCoV-19/Italy/LOM-UniMI-L472/2020\|EPI_ISL_542385\|2 | ···················· | ···················· | ···················· | ···················· | ···················· | ···················· |
| H | 4577 | hCoV-19/Italy/LOM-UniMI-L473/2020\|EPI_ISL_542386\|2 | ···················· | ···················· | ···················· | ···················· | ···················· | ···················· |
| H | 4578 | hCoV-19/Italy/LOM-UniMI-L474/2020\|EPI_ISL_542387\|2 | ···················· | ···················· | ···················· | ···················· | ···················· | ···················· |
| H | 4579 | hCoV-19/Italy/LOM-UniMI-L475/2020\|EPI_ISL_542388\|2 | ···················· | ···················· | ···················· | ···················· | ···················· | ···················· |
| H | 4580 | hCoV-19/Italy/LOM-UniMI-L476/2020\|EPI_ISL_542389\|2 | ···················· | ···················· | ···················· | ···················· | ···················· | ···················· |
| H | 4581 | hCoV-19/Italy/LOM-UniMI-L477/2020\|EPI_ISL_542390\|2 | ···················· | ···················· | ···················· | ···················· | ···················· | ···················· |
| H | 4582 | hCoV-19/Italy/LOM-UniMI-L478/2020\|EPI_ISL_542391\|2 | ···················· | ···················· | ···················· | ···················· | ···················· | ···················· |
| H | 4583 | hCoV-19/Italy/LOM-UniMI-L480/2020\|EPI_ISL_542392\|2 | ···················· | ···················· | ···················· | ···················· | ···················· | ···················· |
| H | 4584 | hCoV-19/Italy/LOM-UniMI-L485/2020\|EPI_ISL_542393\|2 | ···················· | ···················· | ···················· | ···················· | ···················· | ···················· |
| H | 4585 | hCoV-19/Italy/LOM-UniMI-L486/2020\|EPI_ISL_542394\|2 | ···················· | ···················· | ···················· | ···················· | ···················· | ···················· |
| H | 4586 | hCoV-19/Italy/LOM-UniMI-L487/2020\|EPI_ISL_542395\|2 | ···················· | ···················· | ···················· | ···················· | ···················· | ···················· |
| H | 4587 | hCoV-19/Italy/LOM-UniMI-L491/2020\|EPI_ISL_542396\|2 | ···················· | ···················· | ···················· | ···················· | ···················· | ···················· |
| H | 4588 | hCoV-19/Italy/LOM-UniMI-L492/2020\|EPI_ISL_542397\|2 | ···················· | ···················· | ···················· | ···················· | ···················· | ···················· |
| H | 4589 | hCoV-19/Italy/LOM-UniMI-L493/2020\|EPI_ISL_542398\|2 | ···················· | ···················· | ···················· | ···················· | ···················· | ···················· |
| H | 4590 | hCoV-19/Italy/LOM-UniMI-L494/2020\|EPI_ISL_542399\|2 | ···················· | ···················· | ···················· | ···················· | ···················· | ···················· |
| H | 4591 | hCoV-19/USA/TX-HMH-4507/2020\|EPI_ISL_543677\|2020-0 | ···················· | ·············A······ | ···················· | ···················· | ···················· | ···················· |
| H | 4597 | hCoV-19/USA/TX-HMH-4518/2020\|EPI_ISL_543687\|2020-0 | ···················· | ···················· | ···················· | ···················· | ···················· | ···················· |
| H | 4600 | hCoV-19/USA/TX-HMH-4523/2020\|EPI_ISL_543691\|2020-0 | ···················· | ·············A······ | ···················· | ···················· | ···················· | ···················· |
| H | 4601 | hCoV-19/USA/TX-HMH-4526/2020\|EPI_ISL_543692\|2020-0 | ···················· | ···················· | ···················· | ···················· | ···················· | ···················· |
| H | 4604 | hCoV-19/USA/TX-HMH-453/2020\|EPI_ISL_543695\|2020-04 | ···················· | ···················· | ···················· | ···················· | ···················· | ···················· |
| H | 4605 | hCoV-19/USA/TX-HMH-4530/2020\|EPI_ISL_543696\|2020-0 | ···················· | ···················· | ···················· | ···················· | ···················· | ···················· |
| H | 4606 | hCoV-19/USA/TX-HMH-4531/2020\|EPI_ISL_543697\|2020-0 | ···················· | ···················· | ···················· | ···················· | ···················· | ···················· |
| H | 4607 | hCoV-19/USA/TX-HMH-4532/2020\|EPI_ISL_543698\|2020-0 | ···················· | ···················· | ···················· | ···················· | ···················· | ···················· |
| H | 4608 | hCoV-19/USA/TX-HMH-4533/2020\|EPI_ISL_543699\|2020-0 | ···················· | ···················· | ···················· | ···················· | ···················· | ···················· |
| H | 4609 | hCoV-19/England/QEUH-96F18B/2020\|EPI_ISL_530401\|20 | ···················· | ·A·················· | ···················· | ···················· | ···················· | ···················· |
| H | 4610 | hCoV-19/England/QEUH-96E070/2020\|EPI_ISL_530402\|20 | ···················· | ···················· | ···················· | ···················· | ···················· | ···················· |
| H | 4611 | hCoV-19/England/QEUH-96E08F/2020\|EPI_ISL_530403\|20 | ···················· | ·A·················· | ···················· | ···················· | ···················· | ···················· |
| H | 4612 | hCoV-19/England/QEUH-96F1D6/2020\|EPI_ISL_530404\|20 | ···················· | ···················· | ···················· | ···················· | ···················· | ···················· |
| H | 4613 | hCoV-19/England/QEUH-96F5DA/2020\|EPI_ISL_530406\|20 | ···················· | ···················· | ···················· | ···················· | ···················· | ···················· |
| H | 4614 | hCoV-19/England/QEUH-96F84A/2020\|EPI_ISL_530407\|20 | ···················· | ···················· | ···················· | ···················· | ···················· | ···················· |
| H | 4615 | hCoV-19/England/QEUH-96F4A0/2020\|EPI_ISL_530408\|20 | ···················· | ···················· | ···················· | ···················· | ···················· | ···············A···· |
| H | 4616 | hCoV-19/England/QEUH-96F4CE/2020\|EPI_ISL_530409\|20 | ···················· | ···················· | ···················· | ···················· | ···················· | ···················· |
| H | 4617 | hCoV-19/Scotland/QEUH-96E0BC/2020\|EPI_ISL_530410\|2 | ···················· | ···················· | ···················· | ···················· | ···················· | ···················· |
| H | 4618 | hCoV-19/England/QEUH-96E377/2020\|EPI_ISL_530411\|20 | ···················· | ···················· | ···················· | ···················· | ···················· | ···················· |
| H | 4619 | hCoV-19/England/QEUH-96E359/2020\|EPI_ISL_530412\|20 | ···················· | ···················· | ···················· | ···················· | ···················· | ···················· |
| H | 4620 | hCoV-19/England/QEUH-96EFBF/2020\|EPI_ISL_530414\|20 | ···················· | ···················· | ···················· | ···················· | ···················· | ···················· |

| | | | | | | | | |
|---|---|---|---|---|---|---|---|---|
| H | 4621 hCoV-19/England/QEUH-96EED0/2020\|EPI_ISL_530415\|20 | ···················· | ···················· | ···················· | ···················· | ···················· | ···················· | |
| H | 4622 hCoV-19/England/QEUH-96DF1A/2020\|EPI_ISL_530416\|20 | ···················· | ···················· | ···················· | ···················· | ···················· | ···················· | |
| H | 4623 hCoV-19/Northern Ireland/QEUH-96F0BB/2020\|EPI_ISL_ | ········T··········· | ···················· | ···················· | ···················· | ···················· | ···················· | |
| H | 4624 hCoV-19/England/QEUH-96E30E/2020\|EPI_ISL_530418\|20 | ···················· | ···················· | ···················· | ···················· | ···················· | ···················· | |
| H | 4625 hCoV-19/England/QEUH-96F4BF/2020\|EPI_ISL_530419\|20 | ···················· | ···················· | ···················· | ···················· | ···················· | ···················· | |
| H | 4626 hCoV-19/England/QEUH-96F14F/2020\|EPI_ISL_530421\|20 | ···················· | ···················· | ···················· | ···················· | ···················· | ···················· | |
| H | 4627 hCoV-19/England/QEUH-96F1B8/2020\|EPI_ISL_530422\|20 | ···················· | ···················· | ···················· | ···················· | ···················· | ···················· | |
| H | 4628 hCoV-19/England/QEUH-96EF19/2020\|EPI_ISL_530423\|20 | ···················· | ·A·················· | ···················· | ···················· | ···················· | ···A················ | |
| H | 4629 hCoV-19/Northern Ireland/QEUH-96EFEC/2020\|EPI_ISL_ | ···················· | ···················· | ···················· | ···················· | ···················· | ···················· | |
| H | 4630 hCoV-19/England/QEUH-96F130/2020\|EPI_ISL_530425\|20 | ···················· | ·A·····CA··········· | ···················· | ···················· | ···················· | ···················· | 1 |
| H | 4631 hCoV-19/England/QEUH-96EB42/2020\|EPI_ISL_530427\|20 | ···················· | ···················· | ···················· | ···················· | ···················· | ···················· | |
| H | 4632 hCoV-19/Northern Ireland/QEUH-96E887/2020\|EPI_ISL_ | ···················· | ···················· | ···················· | ···················· | ···················· | ···················· | |
| H | 4633 hCoV-19/Scotland/QEUH-96EC9A/2020\|EPI_ISL_530429\|2 | ···················· | ···················· | ···················· | ···················· | ···················· | ···················· | |
| H | 4634 hCoV-19/England/QEUH-96F0AC/2020\|EPI_ISL_530430\|20 | ···················· | ···················· | ···················· | ···················· | ···················· | ···················· | |
| H | 4635 hCoV-19/England/QEUH-96F552/2020\|EPI_ISL_530431\|20 | ···················· | ···················· | ···················· | ···················· | ···················· | ···················· | |
| H | 4636 hCoV-19/England/QEUH-96F710/2020\|EPI_ISL_530433\|20 | ···················· | ···················· | ···················· | ···················· | ···················· | ···················· | |
| H | 4637 hCoV-19/England/QEUH-96DFDE/2020\|EPI_ISL_530434\|20 | ···················· | ···················· | ···················· | ···················· | ···················· | ···················· | |
| H | 4638 hCoV-19/England/QEUH-96E122/2020\|EPI_ISL_530436\|20 | ···················· | ···················· | ···················· | ···················· | ···················· | ···················· | |
| H | 4639 hCoV-19/Northern Ireland/QEUH-96ECE5/2020\|EPI_ISL_ | ···················· | ···················· | ···················· | ···················· | ···················· | ···················· | |
| H | 4640 hCoV-19/Northern Ireland/QEUH-96EB06/2020\|EPI_ISL_ | ···················· | ···················· | ···················· | ···················· | ···················· | ···················· | |
| H | 4641 hCoV-19/England/QEUH-96F5E9/2020\|EPI_ISL_530441\|20 | ···················· | ···················· | ···················· | ···················· | ···················· | ···················· | |
| H | 4642 hCoV-19/England/QEUH-96F103/2020\|EPI_ISL_530442\|20 | ···················· | ·A·················· | ···················· | ···················· | ···················· | ···················· | |
| H | 4643 hCoV-19/England/QEUH-96F8A4/2020\|EPI_ISL_530443\|20 | ···················· | ···················· | ···················· | ···················· | ···················· | ···················· | |
| H | 4644 hCoV-19/Northern Ireland/QEUH-96EEA3/2020\|EPI_ISL_ | ···················· | ···················· | ···················· | ···················· | ···················· | ···················· | |
| H | 4645 hCoV-19/England/QEUH-96EC5E/2020\|EPI_ISL_530445\|20 | ···················· | ···················· | ···················· | ···················· | ···················· | ···················· | |
| H | 4646 hCoV-19/England/QEUH-96EAFA/2020\|EPI_ISL_530447\|20 | ···················· | ···················· | ···················· | ···················· | ···················· | ···················· | |
| H | 4647 hCoV-19/Northern Ireland/QEUH-96E878/2020\|EPI_ISL_ | ···················· | ···················· | ···················· | ···················· | ···················· | ···················· | |
| H | 4648 hCoV-19/England/QEUH-96DFB0/2020\|EPI_ISL_530450\|20 | ···················· | ···················· | ···················· | ···················· | ···················· | ···················· | |
| H | 4649 hCoV-19/Northern Ireland/QEUH-96EA90/2020\|EPI_ISL_ | ···················· | ···················· | ···················· | ···················· | ···················· | ···················· | |
| H | 4650 hCoV-19/Northern Ireland/QEUH-96E4DE/2020\|EPI_ISL_ | ···················· | ···················· | ···················· | ···················· | ···················· | ···················· | |
| H | 4651 hCoV-19/England/QEUH-96F200/2020\|EPI_ISL_530454\|20 | ···················· | ·A·················· | ···················· | ···················· | ···················· | ···················· | |
| H | 4652 hCoV-19/Northern Ireland/QEUH-96EE49/2020\|EPI_ISL_ | ···················· | ···················· | ···················· | ···················· | ···················· | ···················· | |
| H | 4653 hCoV-19/England/QEUH-96F08E/2020\|EPI_ISL_530457\|20 | ···················· | ···················· | ···················· | ···················· | ···················· | ···················· | |
| H | 4654 hCoV-19/England/QEUH-96E052/2020\|EPI_ISL_530458\|20 | ···················· | ···················· | ···················· | ···················· | ···················· | ···················· | |
| H | 4655 hCoV-19/England/QEUH-96F40A/2020\|EPI_ISL_530459\|20 | ···················· | ···················· | ···················· | ···················· | ···················· | ···················· | |
| H | 4656 hCoV-19/England/QEUH-96F72F/2020\|EPI_ISL_530460\|20 | ···················· | ···················· | ···················· | ···················· | ···················· | ···················· | |
| H | 4657 hCoV-19/Northern Ireland/QEUH-96EF37/2020\|EPI_ISL_ | ···················· | ···················· | ···················· | ···················· | ···················· | ···················· | |
| H | 4658 hCoV-19/England/QEUH-96E2D4/2020\|EPI_ISL_530462\|20 | ···················· | ···················· | ···················· | ···················· | ···················· | ···················· | |
| H | 4659 hCoV-19/Northern Ireland/QEUH-96E869/2020\|EPI_ISL_ | ···················· | ···················· | ···················· | ···················· | ···················· | ···················· | |
| H | 4660 hCoV-19/England/QEUH-96E614/2020\|EPI_ISL_530464\|20 | ···················· | ···················· | ···················· | ···················· | ···················· | ···················· | |
| H | 4661 hCoV-19/England/QEUH-96F060/2020\|EPI_ISL_530465\|20 | ···················· | ···················· | ···················· | ···················· | ···················· | ···················· | |
| H | 4662 hCoV-19/England/QEUH-96F3EF/2020\|EPI_ISL_530466\|20 | ···················· | ···················· | ···················· | ···················· | ···················· | ···················· | |
| H | 4663 hCoV-19/England/QEUH-96E7F3/2020\|EPI_ISL_530467\|20 | ···················· | ·A·················· | ···················· | ···················· | ···················· | ···················· | |
| H | 4664 hCoV-19/Scotland/QEUH-96E25C/2020\|EPI_ISL_530468\|2 | ···················· | ···················· | ···················· | ···················· | ···················· | ···················· | |
| H | 4665 hCoV-19/Northern Ireland/QEUH-96EABE/2020\|EPI_ISL_ | ···················· | ···················· | ···················· | ···················· | ···················· | ···················· | |
| H | 4666 hCoV-19/England/QEUH-96F297/2020\|EPI_ISL_530471\|20 | ···················· | ···················· | ···················· | ···················· | ···················· | ···················· | |
| H | 4667 hCoV-19/England/QEUH-96E0F8/2020\|EPI_ISL_530473\|20 | ···················· | ···················· | ···················· | ···················· | ···················· | ···················· | |
| H | 4668 hCoV-19/England/QEUH-96E7E4/2020\|EPI_ISL_530474\|20 | ···················· | ···················· | ···················· | ···················· | ···················· | ···················· | |
| H | 4669 hCoV-19/England/QEUH-96F31C/2020\|EPI_ISL_530476\|20 | ···················· | ···················· | ···················· | ···················· | ···················· | ···················· | |
| H | 4670 hCoV-19/England/QEUH-96DFFC/2020\|EPI_ISL_530477\|20 | ···················· | ·A·················· | ···················· | ···················· | ···················· | ···A················ | |
| H | 4671 hCoV-19/England/QEUH-96E5BD/2020\|EPI_ISL_530478\|20 | ···················· | ···················· | ···················· | ···················· | ···················· | ···················· | |
| H | 4672 hCoV-19/Scotland/QEUH-96EC12/2020\|EPI_ISL_530480\|2 | ···················· | ···················· | ···················· | ···················· | ···················· | ···················· | |
| H | 4673 hCoV-19/England/QEUH-96EEEF/2020\|EPI_ISL_530483\|20 | ···················· | ···················· | ···················· | ···················· | ···················· | ···················· | |
| H | 4674 hCoV-19/England/QEUH-96F5AD/2020\|EPI_ISL_530484\|20 | ···················· | ···················· | ···················· | ···················· | ···················· | ···················· | |
| H | 4675 hCoV-19/England/QEUH-96EDA6/2020\|EPI_ISL_530486\|20 | ···················· | ·A·················· | ···················· | ···················· | ···················· | ···················· | |
| H | 4676 hCoV-19/England/QEUH-96EC21/2020\|EPI_ISL_530487\|20 | ···················· | ·A·················· | ···················· | ···················· | ···················· | ···················· | |
| H | 4677 hCoV-19/England/QEUH-962935/2020\|EPI_ISL_531733\|20 | ···················· | ···················· | ···················· | ···················· | ···················· | ···················· | |
| H | 4678 hCoV-19/England/QEUH-962610/2020\|EPI_ISL_531739\|20 | ···················· | ···················· | ···················· | ···················· | ···················· | ···················· | |
| H | 4679 hCoV-19/Wales/QEUH-963318/2020\|EPI_ISL_531741\|2020 | ···················· | ···················· | ···················· | ···················· | ···················· | ···················· | |
| H | 4680 hCoV-19/Scotland/QEUH-96357C/2020\|EPI_ISL_531743\|2 | ···················· | ···················· | ···················· | ···················· | ···················· | ···················· | |
| H | 4681 hCoV-19/England/QEUH-9637FE/2020\|EPI_ISL_531744\|20 | ···················· | ·A·················· | ···················· | ···················· | ···················· | ···················· | |
| H | 4682 hCoV-19/England/QEUH-963855/2020\|EPI_ISL_531745\|20 | ···················· | ·A·················· | ···················· | ···················· | ···················· | ···A················ | |
| H | 4683 hCoV-19/England/QEUH-9620D6/2020\|EPI_ISL_531746\|20 | ···················· | ···················· | ···················· | ···················· | ···················· | ···················· | |
| H | 4684 hCoV-19/England/QEUH-962470/2020\|EPI_ISL_531748\|20 | ···················· | ···················· | ···················· | ···················· | ···················· | ···················· | |
| H | 4685 hCoV-19/England/QEUH-962416/2020\|EPI_ISL_531750\|20 | ···················· | ···················· | ···················· | ···················· | ···················· | ···················· | |
| H | 4686 hCoV-19/Northern Ireland/QEUH-9620F4/2020\|EPI_ISL_ | ···················· | ···················· | ···················· | ···················· | ···················· | ···················· | |
| H | 4687 hCoV-19/England/QEUH-962DEE/2020\|EPI_ISL_531752\|20 | ···················· | ···················· | ···················· | ···················· | ···················· | ···················· | |
| H | 4688 hCoV-19/Scotland/QEUH-9636E2/2020\|EPI_ISL_531754\|2 | ···················· | ···················· | ···················· | ···················· | ···················· | ···················· | |
| H | 4689 hCoV-19/Scotland/QEUH-96372B/2020\|EPI_ISL_531755\|2 | ···················· | ···················· | ···················· | ···················· | ···················· | ···················· | |
| H | 4690 hCoV-19/England/QEUH-96370D/2020\|EPI_ISL_531756\|20 | ···················· | ···················· | ···················· | ···················· | ···················· | ···················· | |
| H | 4691 hCoV-19/England/QEUH-9636F1/2020\|EPI_ISL_531757\|20 | ···················· | ···················· | ···················· | ···················· | ···················· | ···················· | |
| H | 4692 hCoV-19/England/QEUH-96373A/2020\|EPI_ISL_531759\|20 | ···················· | ·A·················· | ···················· | ···················· | ···················· | ···················· | |
| H | 4693 hCoV-19/England/QEUH-9628FC/2020\|EPI_ISL_531760\|20 | ···················· | ···················· | ···················· | ···················· | ···················· | ···················· | |
| H | 4694 hCoV-19/Northern Ireland/QEUH-962249/2020\|EPI_ISL_ | ···················· | ···················· | ···················· | ···················· | ···················· | ···················· | |
| H | 4695 hCoV-19/England/QEUH-9638EC/2020\|EPI_ISL_531762\|20 | ···················· | ···················· | ···················· | ···················· | ···················· | ···················· | |
| H | 4696 hCoV-19/Northern Ireland/QEUH-9634CA/2020\|EPI_ISL_ | ···················· | ···················· | ···················· | ···················· | ···················· | ···················· | |
| H | 4697 hCoV-19/England/QEUH-962C5A/2020\|EPI_ISL_531764\|20 | ···················· | ···················· | ···················· | ···················· | ···················· | ···················· | |
| H | 4698 hCoV-19/England/QEUH-96365B/2020\|EPI_ISL_531766\|20 | ···················· | ·A·················· | ···················· | ···················· | ···················· | ···A················ | |
| H | 4699 hCoV-19/Northern Ireland/QEUH-9622D0/2020\|EPI_ISL_ | ···················· | ···················· | ···················· | ···················· | ···················· | ···················· | |
| H | 4700 hCoV-19/Scotland/QEUH-962AAB/2020\|EPI_ISL_531768\|2 | ···················· | ···················· | ···················· | ···················· | ···················· | ···················· | |

| | | | | | | | |
|---|---|---|---|---|---|---|---|
| H | 4701 hCoV-19/Northern Ireland/QEUH-962267/2020\|EPI_ISL_ | ···················· | ···················· | ····················· | ···················· | ···················· | ···················· |
| H | 4702 hCoV-19/England/QEUH-963512/2020\|EPI_ISL_531770\|20 | ···················· | ···················· | ····················· | ···················· | ···················· | ···················· |
| H | 4703 hCoV-19/England/QEUH-962FAC/2020\|EPI_ISL_531771\|20 | ···················· | ···················· | ····················· | ···················· | ···················· | ···················· |
| H | 4704 hCoV-19/England/QEUH-9632FD/2020\|EPI_ISL_531772\|20 | ···················· | ···················· | ····················· | ···················· | ···················· | ···················· |
| H | 4705 hCoV-19/Northern Ireland/QEUH-9630A8/2020\|EPI_ISL_ | ···················· | ···················· | ····················· | ···················· | ···················· | ···················· |
| H | 4706 hCoV-19/England/QEUH-962F51/2020\|EPI_ISL_531774\|20 | ···················· | ···················· | ····················· | ···················· | ···················· | ···················· |
| H | 4707 hCoV-19/Wales/QEUH-963679/2020\|EPI_ISL_531776\|2020 | ···················· | ···················· | ····················· | ···················· | ···················· | ···················· |
| H | 4708 hCoV-19/England/QEUH-9636A6/2020\|EPI_ISL_531777\|20 | ···················· | ···················· | ····················· | ···················· | ···················· | ···················· |
| H | 4709 hCoV-19/England/QEUH-963882/2020\|EPI_ISL_531778\|20 | ···················· | ·A·················· | ····················· | ···················· | ···················· | ···A················ |
| H | 4710 hCoV-19/England/QEUH-962425/2020\|EPI_ISL_531780\|20 | ···················· | ···················· | ····················· | ···················· | ····A··············· | ···················· |
| H | 4711 hCoV-19/England/QEUH-962926/2020\|EPI_ISL_531781\|20 | ···················· | ···················· | ····················· | ···················· | ···················· | ···················· |
| H | 4712 hCoV-19/England/QEUH-963433/2020\|EPI_ISL_531782\|20 | ···················· | ···················· | ····················· | ···················· | ···················· | ···················· |
| H | 4713 hCoV-19/England/QEUH-96272C/2020\|EPI_ISL_531783\|20 | ···················· | ···················· | ····················· | ···················· | ···················· | ···················· |
| H | 4714 hCoV-19/Northern Ireland/QEUH-962F24/2020\|EPI_ISL_ | ···················· | ···················· | ····················· | ···················· | ···················· | ···················· |
| H | 4715 hCoV-19/England/QEUH-963372/2020\|EPI_ISL_531786\|20 | ···················· | ·A·················· | ····················· | ···················· | ···················· | ···A················ |
| H | 4716 hCoV-19/England/QEUH-963785/2020\|EPI_ISL_531787\|20 | ···················· | ···················· | ····················· | ···················· | ···················· | ···················· |
| H | 4717 hCoV-19/England/QEUH-963275/2020\|EPI_ISL_531790\|20 | ···················· | ···················· | ····················· | ···················· | ···················· | ···················· |
| H | 4718 hCoV-19/Northern Ireland/QEUH-962030/2020\|EPI_ISL_ | ···················· | ···················· | ····················· | ···················· | ···················· | ···················· |
| H | 4719 hCoV-19/England/QEUH-9635D6/2020\|EPI_ISL_531793\|20 | ···················· | ···················· | ····················· | ···················· | ···················· | ···················· |
| H | 4720 hCoV-19/Northern Ireland/QEUH-9621C4/2020\|EPI_ISL_ | ···················· | ···················· | ····················· | ···················· | ···················· | ···················· |
| H | 4721 hCoV-19/Northern Ireland/QEUH-962434/2020\|EPI_ISL_ | ···················· | ···················· | ····················· | ···················· | ···················· | ···················· |
| H | 4722 hCoV-19/Wales/QEUH-96371C/2020\|EPI_ISL_531796\|2020 | ···················· | ···················· | ····················· | ···················· | ···················· | ···················· |
| H | 4723 hCoV-19/Northern Ireland/QEUH-96322A/2020\|EPI_ISL_ | ···················· | ···················· | ····················· | ···················· | ···················· | ···················· |
| H | 4724 hCoV-19/England/QEUH-962A8D/2020\|EPI_ISL_531798\|20 | ···················· | ···················· | ····················· | ···················· | ···················· | ···················· |
| H | 4725 hCoV-19/England/QEUH-961F43/2020\|EPI_ISL_531799\|20 | ···················· | ···················· | ····················· | ···················· | ···················· | ···················· |
| H | 4727 hCoV-19/Wales/PHWC-16A4E9/2020\|EPI_ISL_535261\|2020 | ···················· | ···················· | ····················· | ···················· | ···················· | ···················· |
| H | 4728 hCoV-19/USA/NM-NMDOH-2020099249/2020\|EPI_ISL_53526 | ···················· | ···················· | ····················· | ···················· | ···················· | ···················· |
| H | 4729 hCoV-19/USA/NM-NMDOH-2020099251/2020\|EPI_ISL_53526 | ···················· | ···················· | ····················· | ···················· | ···················· | ···················· |
| H | 4730 hCoV-19/USA/NM-NMDOH-2020099255/2020\|EPI_ISL_53526 | ···················· | ···················· | ····················· | ···················· | ···················· | ···················· |
| H | 4731 hCoV-19/USA/NM-NMDOH-2020099258/2020\|EPI_ISL_53526 | ···················· | ···················· | ····················· | ···················· | ···················· | ···················· |
| H | 4732 hCoV-19/USA/NM-NMDOH-2020126501/2020\|EPI_ISL_53526 | ···················· | ···················· | ····················· | ···················· | ···················· | ···················· |
| H | 4733 hCoV-19/USA/NM-NMDOH-2020126502/2020\|EPI_ISL_53527 | ···················· | ···················· | ····················· | ···················· | ···················· | ···················· |
| H | 4734 hCoV-19/USA/NM-NMDOH-2020126537/2020\|EPI_ISL_53527 | ···················· | ···················· | ····················· | ···················· | ···················· | ···················· |
| H | 4735 hCoV-19/USA/NM-NMDOH-2020126547/2020\|EPI_ISL_53527 | ···················· | ···················· | ····················· | ···················· | ···················· | ···················· |
| H | 4736 hCoV-19/USA/NM-NMDOH-2020126554/2020\|EPI_ISL_53527 | ···················· | ···················· | ····················· | ···················· | ···················· | ···················· |
| H | 4737 hCoV-19/USA/NM-NMDOH-2020126556/2020\|EPI_ISL_53527 | ···················· | ···················· | ····················· | ···················· | ···················· | ···················· |
| H | 4738 hCoV-19/USA/NM-NMDOH-2020126568/2020\|EPI_ISL_53527 | ···················· | ···················· | ····················· | ···················· | ···················· | ···················· |
| H | 4739 hCoV-19/USA/NM-NMDOH-2020134982/2020\|EPI_ISL_53527 | ···················· | ···················· | ····················· | ···················· | ···················· | ···················· |
| H | 4740 hCoV-19/USA/NM-NMDOH-2020134983/2020\|EPI_ISL_53527 | ···················· | ···················· | ····················· | ···················· | ···················· | ···················· |
| H | 4741 hCoV-19/USA/NM-NMDOH-2020134985/2020\|EPI_ISL_53528 | ···················· | ···················· | ····················· | ···················· | ···················· | ···················· |
| H | 4742 hCoV-19/USA/NM-NMDOH-2020134989/2020\|EPI_ISL_53528 | ···················· | ···················· | ····················· | ···················· | ···················· | ···················· |
| H | 4743 hCoV-19/USA/NM-NMDOH-2020134990/2020\|EPI_ISL_53528 | ···················· | ···················· | ····················· | ···················· | ···················· | ···················· |
| H | 4744 hCoV-19/USA/NM-NMDOH-2020134991/2020\|EPI_ISL_53528 | ···················· | ···················· | ····················· | ···················· | ···················· | ···················· |
| H | 4745 hCoV-19/USA/NM-NMDOH-2020134997/2020\|EPI_ISL_53528 | ···················· | ···················· | ····················· | ···················· | ···················· | ···················· |
| H | 4746 hCoV-19/USA/NM-NMDOH-2020134998/2020\|EPI_ISL_53528 | ···················· | ···················· | ····················· | ···················· | ···················· | ···················· |
| H | 4747 hCoV-19/USA/NM-NMDOH-2020134999/2020\|EPI_ISL_53528 | ···················· | ···················· | ····················· | ···················· | ···················· | ···················· |
| H | 4748 hCoV-19/USA/NM-NMDOH-2020135000/2020\|EPI_ISL_53528 | ···················· | ···················· | ····················· | ···················· | ···················· | ···················· |
| H | 4749 hCoV-19/USA/WI-UW-1217/2020\|EPI_ISL_536590\|2020-08 | ···················· | ···················· | ····················· | ···················· | ···················· | ···················· |
| H | 4750 hCoV-19/USA/WI-UW-1218/2020\|EPI_ISL_536591\|2020-08 | ···················· | ···················· | ····················· | ···················· | ···················· | ···················· |
| H | 4752 hCoV-19/USA/WI-UW-1220/2020\|EPI_ISL_536593\|2020-08 | ···················· | ···················· | ····················· | ···················· | ···················· | ···················· |
| H | 4753 hCoV-19/USA/WI-UW-1221/2020\|EPI_ISL_536594\|2020-08 | ···················· | ···················· | ····················· | ···················· | ···················· | ···················· |
| H | 4754 hCoV-19/USA/WI-UW-1222/2020\|EPI_ISL_536595\|2020-08 | ···················· | ···················· | ····················· | ···················· | ···················· | ···················· |
| H | 4755 hCoV-19/USA/WI-UW-1223/2020\|EPI_ISL_536596\|2020-08 | ···················· | ···················· | ····················· | ···················· | ···················· | ···················· |
| H | 4756 hCoV-19/USA/WI-UW-1224/2020\|EPI_ISL_536597\|2020-08 | ···················· | ···················· | ····················· | ···················· | ···················· | ···················· |
| H | 4757 hCoV-19/USA/WI-UW-1225/2020\|EPI_ISL_536598\|2020-08 | ···················· | ·A·················· | ····················· | ···················· | ···················· | ···················· |
| H | 4758 hCoV-19/USA/WI-UW-1226/2020\|EPI_ISL_536599\|2020-08 | ···················· | ···················· | ····················· | ···················· | ···················· | ···················· |
| H | 4759 hCoV-19/Italy/LOM-UniMI-L53/2020\|EPI_ISL_542400\|20 | ···················· | ···················· | ····················· | ···················· | ···················· | ···················· |
| H | 4760 hCoV-19/Italy/LOM-UniMI-L54/2020\|EPI_ISL_542401\|20 | ···················· | ···················· | ····················· | ···················· | ···················· | ···················· |
| H | 4761 hCoV-19/Italy/LOM-UniMI-L55/2020\|EPI_ISL_542402\|20 | ···················· | ···················· | ····················· | ···················· | ···················· | ···················· |
| H | 4762 hCoV-19/Italy/LOM-UniMI-L56/2020\|EPI_ISL_542403\|20 | ···················· | ···················· | ····················· | ···················· | ···················· | ···················· |
| H | 4763 hCoV-19/Italy/LOM-UniMI-L57/2020\|EPI_ISL_542404\|20 | ···················· | ···················· | ····················· | ···················· | ···················· | ···················· |
| H | 4764 hCoV-19/Italy/LOM-UniMI-L58/2020\|EPI_ISL_542405\|20 | ···················· | ···················· | ····················· | ···················· | ···················· | ···················· |
| H | 4765 hCoV-19/Italy/LOM-UniMI-L60/2020\|EPI_ISL_542406\|20 | ···················· | ···················· | ····················· | ···················· | ···················· | ···················· |
| H | 4766 hCoV-19/Italy/LOM-UniMI-L61/2020\|EPI_ISL_542407\|20 | ···················· | ···················· | ····················· | ···················· | ···················· | ···················· |
| H | 4767 hCoV-19/Italy/LOM-UniMI-L62/2020\|EPI_ISL_542408\|20 | ···················· | ···················· | ····················· | ···················· | ···················· | ···················· |
| H | 4768 hCoV-19/Italy/LOM-UniMI-L63/2020\|EPI_ISL_542409\|20 | ···················· | ···················· | ····················· | ···················· | ···················· | ···················· |
| H | 4769 hCoV-19/Italy/LOM-UniMI-L64/2020\|EPI_ISL_542410\|20 | ···················· | ···················· | ····················· | ···················· | ···················· | ···················· |
| H | 4770 hCoV-19/Italy/LOM-UniMI-L65/2020\|EPI_ISL_542411\|20 | ···················· | ···················· | ····················· | ···················· | ···················· | ···················· |
| H | 4771 hCoV-19/Italy/LOM-UniMI-L66/2020\|EPI_ISL_542412\|20 | ···················· | ···················· | ····················· | ···················· | ···················· | ···················· |
| H | 4772 hCoV-19/Italy/LOM-UniMI-L67/2020\|EPI_ISL_542413\|20 | ···················· | ···················· | ····················· | ···················· | ···················· | ···················· |
| H | 4773 hCoV-19/Italy/LOM-UniMI-L68/2020\|EPI_ISL_542414\|20 | ···················· | ···················· | ····················· | ···················· | ···················· | ···················· |
| H | 4774 hCoV-19/Italy/LOM-UniMI-L69/2020\|EPI_ISL_542415\|20 | ···················· | ···················· | ····················· | ···················· | ···················· | ···················· |
| H | 4775 hCoV-19/Italy/LOM-UniMI-L70/2020\|EPI_ISL_542416\|20 | ···················· | ···················· | ····················· | ···················· | ···················· | ···················· |
| H | 4776 hCoV-19/Italy/LOM-UniMI-L71/2020\|EPI_ISL_542417\|20 | ···················· | ···················· | ····················· | ···················· | ···················· | ···················· |
| H | 4777 hCoV-19/Italy/LOM-UniMI-L73/2020\|EPI_ISL_542418\|20 | ···················· | ···················· | ····················· | ···················· | ···················· | ···················· |
| H | 4778 hCoV-19/Italy/LOM-UniMI-L74/2020\|EPI_ISL_542419\|20 | ···················· | ···················· | ····················· | ···················· | ···················· | ···················· |
| H | 4779 hCoV-19/Italy/LOM-UniMI-L75/2020\|EPI_ISL_542420\|20 | ···················· | ···················· | ····················· | ···················· | ···················· | ···················· |
| H | 4780 hCoV-19/Italy/LOM-UniMI-L76/2020\|EPI_ISL_542421\|20 | ···················· | ···················· | ····················· | ···················· | ···················· | ···················· |
| H | 4781 hCoV-19/Italy/LOM-UniMI-L77/2020\|EPI_ISL_542422\|20 | ···················· | ···················· | ····················· | ···················· | ···················· | ···················· |
| H | 4782 hCoV-19/Italy/LOM-UniMI-L78/2020\|EPI_ISL_542423\|20 | ···················· | ···················· | ····················· | ···················· | ···················· | ···················· |

| | | | | | | | | | |
|---|---|---|---|---|---|---|---|---|---|
| H | 4783 | hCoV-19/Italy/LOM-UniMI-L79/2020\|EPI_ISL_542424\|20 | ···················· | ···················· | ···················· | ···················· | ···················· | ···················· | |
| H | 4784 | hCoV-19/Italy/LOM-UniMI-L80/2020\|EPI_ISL_542425\|20 | ···················· | ···················· | ···················· | ···················· | ···················· | ···················· | |
| H | 4785 | hCoV-19/Italy/LOM-UniMI-L81/2020\|EPI_ISL_542426\|20 | ···················· | ···················· | ···················· | ···················· | ···················· | ···················· | |
| H | 4786 | hCoV-19/Italy/LOM-UniMI-L83/2020\|EPI_ISL_542427\|20 | ···················· | ···················· | ···················· | ···················· | ···················· | ···················· | |
| H | 4787 | hCoV-19/Italy/LOM-UniMI-L84/2020\|EPI_ISL_542428\|20 | ···················· | ···················· | ···················· | ···················· | ···················· | ···················· | |
| H | 4788 | hCoV-19/Italy/LOM-UniMI-L85/2020\|EPI_ISL_542429\|20 | ···················· | ···················· | ···················· | ···················· | ···················· | ···················· | |
| H | 4789 | hCoV-19/Italy/LOM-UniMI-L86/2020\|EPI_ISL_542430\|20 | ···················· | ···················· | ···················· | ···················· | ···················· | ···················· | |
| H | 4790 | hCoV-19/Italy/LOM-UniMI-L87/2020\|EPI_ISL_542431\|20 | ···················· | ···················· | ···················· | ···················· | ···················· | ···················· | |
| H | 4791 | hCoV-19/Italy/LOM-UniMI-L88/2020\|EPI_ISL_542432\|20 | ···················· | ···················· | ···················· | ···················· | ···················· | ···················· | |
| H | 4792 | hCoV-19/Italy/LOM-UniMI-L89/2020\|EPI_ISL_542433\|20 | ···················· | ···················· | ···················· | ···················· | ···················· | ···················· | |
| H | 4793 | hCoV-19/Italy/LOM-UniMI-L90/2020\|EPI_ISL_542434\|20 | ···················· | ···················· | ···················· | ···················· | ···················· | ···················· | |
| H | 4794 | hCoV-19/Italy/LOM-UniMI-L91/2020\|EPI_ISL_542435\|20 | ···················· | ···················· | ···················· | ···················· | ···················· | ···················· | |
| H | 4795 | hCoV-19/Italy/LOM-UniMI-L92/2020\|EPI_ISL_542436\|20 | ···················· | ···················· | ···················· | ···················· | ···················· | ···················· | |
| H | 4796 | hCoV-19/Italy/LOM-UniMI-L93/2020\|EPI_ISL_542437\|20 | ···················· | ···················· | ···················· | ···················· | ···················· | ···················· | |
| H | 4797 | hCoV-19/Italy/LOM-UniMI-L94/2020\|EPI_ISL_542438\|20 | ···················· | ···················· | ···················· | ···················· | ···················· | ···················· | |
| H | 4798 | hCoV-19/Italy/LOM-UniMI-L95/2020\|EPI_ISL_542439\|20 | ···················· | ···················· | ···················· | ···················· | ···················· | ···················· | |
| H | 4799 | hCoV-19/Italy/LOM-UniMI-L96/2020\|EPI_ISL_542440\|20 | ···················· | ···················· | ···················· | ···················· | ···················· | ···················· | |
| H | 4800 | hCoV-19/Italy/LOM-UniMI-L97/2020\|EPI_ISL_542441\|20 | ···················· | ···················· | ···················· | ···················· | ···················· | ···················· | |
| H | 4801 | hCoV-19/Italy/LOM-UniMI-L98/2020\|EPI_ISL_542442\|20 | ···················· | ···················· | ···················· | ···················· | ···················· | ···················· | |
| H | 4802 | hCoV-19/Italy/LOM-UniMI-L99/2020\|EPI_ISL_542443\|20 | ···················· | ···················· | ···················· | ···················· | ···················· | ···················· | |
| H | 4803 | hCoV-19/USA/TX-DSHS-0404/2020\|EPI_ISL_542464\|2020- | ···················· | ···················· | ···················· | ···················· | ···················· | ···················· | |
| H | 4804 | hCoV-19/USA/TX-HMH-4547/2020\|EPI_ISL_543710\|2020-0 | ···················· | ···················· | ···················· | ···················· | ···················· | ···················· | |
| H | 4805 | hCoV-19/USA/TX-HMH-4551/2020\|EPI_ISL_543711\|2020-0 | ···················· | ···················· | ···················· | ···················· | ···················· | ···················· | |
| H | 4806 | hCoV-19/USA/TX-HMH-4552/2020\|EPI_ISL_543712\|2020-0 | ···················· | ···················· | ···················· | ···················· | ···················· | ···················· | |
| H | 4807 | hCoV-19/USA/TX-HMH-4556/2020\|EPI_ISL_543715\|2020-0 | ···················· | ·············A······ | ···················· | ···················· | ···················· | ···················· | |
| H | 4808 | hCoV-19/USA/TX-HMH-4559/2020\|EPI_ISL_543717\|2020-0 | ···················· | ···················· | ···················· | ···················· | ···················· | ···················· | |
| H | 4813 | hCoV-19/USA/TX-HMH-4569/2020\|EPI_ISL_543723\|2020-0 | ···················· | ···················· | ···················· | ···················· | ···················· | ···················· | |
| H | 4814 | hCoV-19/USA/TX-HMH-457/2020\|EPI_ISL_543724\|2020-04 | ···················· | ···················· | ···················· | ···················· | ···················· | ···················· | |
| H | 4815 | hCoV-19/USA/TX-HMH-4571/2020\|EPI_ISL_543725\|2020-0 | ···················· | ·A·················· | ···················· | ···················· | ···················· | ···················· | |
| H | 4819 | hCoV-19/USA/TX-HMH-4579/2020\|EPI_ISL_543729\|2020-0 | ···················· | ···················· | ···················· | ···················· | ···················· | ···················· | |
| H | 4820 | hCoV-19/USA/TX-HMH-4580/2020\|EPI_ISL_543730\|2020-0 | ···················· | ···················· | ···················· | ···················· | ···················· | ···················· | |
| H | 4822 | hCoV-19/USA/TX-HMH-4582/2020\|EPI_ISL_543732\|2020-0 | ···················· | ···················· | ···················· | ···················· | ···················· | ···················· | |
| H | 4824 | hCoV-19/USA/TX-HMH-4584/2020\|EPI_ISL_543734\|2020-0 | ···················· | ···················· | ···················· | ···················· | ···················· | ···················· | |
| H | 4825 | hCoV-19/USA/TX-HMH-4585/2020\|EPI_ISL_543735\|2020-0 | ···················· | ···················· | ···················· | ···················· | ···················· | ···················· | |
| H | 4827 | hCoV-19/USA/TX-HMH-4587/2020\|EPI_ISL_543737\|2020-0 | ···················· | ···················· | ···················· | ···················· | ···················· | ···················· | |
| H | 4830 | hCoV-19/USA/TX-HMH-4593/2020\|EPI_ISL_543740\|2020-0 | ···················· | ···················· | ···················· | ···················· | ···················· | NN·················· | |
| H | 4833 | hCoV-19/USA/TX-HMH-4598/2020\|EPI_ISL_543744\|2020-0 | ···················· | ···················· | ···················· | ···················· | ···················· | ···················· | |
| H | 4838 | hCoV-19/USA/TX-HMH-461/2020\|EPI_ISL_543753\|2020-04 | ···················· | ···················· | ···················· | ···················· | ···················· | ···················· | |
| H | 4839 | hCoV-19/USA/TX-HMH-4615/2020\|EPI_ISL_543756\|2020-0 | ···················· | ···················· | ···················· | ···················· | ···················· | ···················· | |
| H | 4840 | hCoV-19/USA/TX-HMH-4616/2020\|EPI_ISL_543757\|2020-0 | ···················· | ···················· | ···················· | ···················· | ···················· | ···················· | |
| H | 4842 | hCoV-19/USA/TX-HMH-4618/2020\|EPI_ISL_543759\|2020-0 | ···················· | ···················· | ···················· | ···················· | ···················· | ···················· | |
| H | 4843 | hCoV-19/USA/TX-HMH-4619/2020\|EPI_ISL_543760\|2020-0 | ···················· | ···················· | ···················· | ···················· | ···················· | N··················· | |
| H | 4845 | hCoV-19/USA/TX-HMH-4623/2020\|EPI_ISL_543762\|2020-0 | ···················· | ···················· | ···················· | ···················· | ···················· | ··A················ | |
| H | 4846 | hCoV-19/USA/TX-HMH-4624/2020\|EPI_ISL_543763\|2020-0 | ···················· | ···················· | ···················· | ···················· | ···················· | ···················· | |
| H | 4849 | hCoV-19/USA/TX-HMH-463/2020\|EPI_ISL_543767\|2020-04 | ···················· | ···················· | ···················· | ···················· | ···················· | ···················· | |
| H | 4850 | hCoV-19/USA/TX-HMH-4630/2020\|EPI_ISL_543768\|2020-0 | ···················· | ···················· | ···················· | ···················· | ···················· | ···················· | |
| H | 4852 | hCoV-19/USA/TX-HMH-4632/2020\|EPI_ISL_543770\|2020-0 | ···················· | ···················· | ···················· | ···················· | ···················· | NN·················· | |
| H | 4854 | hCoV-19/USA/TX-HMH-4636/2020\|EPI_ISL_543773\|2020-0 | ···················· | ···················· | ···················· | ···················· | ···················· | ···················· | |
| H | 4855 | hCoV-19/USA/TX-HMH-4637/2020\|EPI_ISL_543774\|2020-0 | ···················· | ···················· | ···················· | ···················· | ··············N····· | ·············N······ | |
| H | 4856 | hCoV-19/USA/TX-HMH-4638/2020\|EPI_ISL_543775\|2020-0 | ···················· | ···················· | ···················· | ···················· | ···················· | NN···········N······ | 1 |
| H | 4857 | hCoV-19/USA/TX-HMH-464/2020\|EPI_ISL_543776\|2020-04 | ···················· | ···················· | ···················· | ···················· | ···················· | ···················· | |
| H | 4859 | hCoV-19/USA/TX-HMH-4652/2020\|EPI_ISL_543781\|2020-0 | ···················· | ···················· | ···················· | ···················· | ···················· | ···················· | |
| H | 4860 | hCoV-19/USA/TX-HMH-4654/2020\|EPI_ISL_543782\|2020-0 | ···················· | ···················· | ···················· | ···················· | ···················· | N··················· | |
| H | 4863 | hCoV-19/USA/TX-HMH-466/2020\|EPI_ISL_543786\|2020-04 | ···················· | ···················· | ···················· | ···················· | ···················· | ···················· | |
| H | 4865 | hCoV-19/Australia/QLD1005/2020\|EPI_ISL_530246\|2020 | ···················· | ···················· | ···················· | ···················· | ···················· | ···················· | |
| H | 4866 | hCoV-19/Australia/QLD1006/2020\|EPI_ISL_530247\|2020 | ···················· | ···················· | ···················· | ···················· | ···················· | ···················· | |
| H | 4867 | hCoV-19/Australia/QLD1007/2020\|EPI_ISL_530248\|2020 | ···················· | ···················· | ···················· | ···················· | ···················· | ···················· | |
| H | 4868 | hCoV-19/Australia/QLD1008/2020\|EPI_ISL_530249\|2020 | ···················· | ···················· | ···················· | ···················· | ···················· | ···················· | |
| H | 4869 | hCoV-19/Australia/QLD990/2020\|EPI_ISL_530250\|2020- | ···················· | ···················· | ···················· | ···················· | ···················· | ···················· | |
| H | 4870 | hCoV-19/Australia/QLD991/2020\|EPI_ISL_530251\|2020- | ···················· | ···················· | ···················· | ···················· | ···················· | ···················· | |
| H | 4871 | hCoV-19/Australia/QLD992/2020\|EPI_ISL_530252\|2020- | ···················· | ···················· | ···················· | ···················· | ···················· | ···················· | |
| H | 4872 | hCoV-19/Australia/QLD993/2020\|EPI_ISL_530253\|2020- | ···················· | ···················· | ···················· | ···················· | ···················· | ···················· | |
| H | 4873 | hCoV-19/Australia/QLD994/2020\|EPI_ISL_530254\|2020- | ···················· | ···················· | ···················· | ···················· | ···················· | ···················· | |
| H | 4874 | hCoV-19/Australia/QLD996/2020\|EPI_ISL_530255\|2020- | ···················· | ···················· | ···················· | ···················· | ···················· | ···················· | |
| H | 4875 | hCoV-19/Australia/QLD998/2020\|EPI_ISL_530256\|2020- | ···················· | ···················· | ···················· | ···················· | ···················· | ···················· | |
| H | 4876 | hCoV-19/Australia/QLD999/2020\|EPI_ISL_530257\|2020- | ···················· | ···················· | ···················· | ···················· | ···················· | ···················· | |
| H | 4877 | hCoV-19/Australia/QLD1000/2020\|EPI_ISL_530258\|2020 | ···················· | ···················· | ···················· | ···················· | ···················· | ···················· | |
| H | 4878 | hCoV-19/Australia/QLD1001/2020\|EPI_ISL_530259\|2020 | ···················· | ···················· | ···················· | ···················· | ···················· | ···················· | |
| H | 4879 | hCoV-19/Australia/QLD980/2020\|EPI_ISL_530260\|2020- | ···················· | ···················· | ···················· | ···················· | ···················· | ···················· | |
| H | 4880 | hCoV-19/Australia/QLD981/2020\|EPI_ISL_530261\|2020- | ···················· | ···················· | ···················· | ···················· | ···················· | ···················· | |
| H | 4881 | hCoV-19/Australia/QLD982/2020\|EPI_ISL_530262\|2020- | ···················· | ···················· | ···················· | ···················· | ···················· | ···················· | |
| H | 4882 | hCoV-19/Australia/QLD983/2020\|EPI_ISL_530263\|2020- | ···················· | ···················· | ···················· | ···················· | ···················· | ···················· | |
| H | 4883 | hCoV-19/Australia/QLD984/2020\|EPI_ISL_530264\|2020- | ···················· | ···················· | ···················· | ···················· | ···················· | ···················· | |
| H | 4884 | hCoV-19/Australia/QLD985/2020\|EPI_ISL_530265\|2020- | ···················· | ···················· | ···················· | ···················· | ···················· | ···················· | |
| H | 4885 | hCoV-19/Australia/QLD986/2020\|EPI_ISL_530266\|2020- | ···················· | ···················· | ···················· | ···················· | ···················· | ···················· | |
| H | 4886 | hCoV-19/Australia/QLD987/2020\|EPI_ISL_530267\|2020- | ···················· | ···················· | ···················· | ···················· | ···················· | ···················· | |
| H | 4887 | hCoV-19/Australia/QLD988/2020\|EPI_ISL_530268\|2020- | ···················· | ···················· | ···················· | ···················· | ···················· | ···················· | |
| H | 4888 | hCoV-19/Australia/QLD1009/2020\|EPI_ISL_530269\|2020 | ···················· | ···················· | ···················· | ···················· | ···················· | ···················· | |
| H | 4889 | hCoV-19/Australia/QLD1010/2020\|EPI_ISL_530270\|2020 | ···················· | ···················· | ···················· | ···················· | ···················· | ···················· | |
| H | 4890 | hCoV-19/Australia/QLD1011/2020\|EPI_ISL_530271\|2020 | ···················· | ···················· | ···················· | ···················· | ···················· | ···················· | |

H 4891 hCoV-19/Australia/QLD1012/2020|EPI_ISL_530272|2020 ···················· ···················· ···················· ···················· ···················· ····················
H 4892 hCoV-19/Australia/QLD1013/2020|EPI_ISL_530273|2020 ···················· ···················· ···················· ···················· ···················· ····················
H 4893 hCoV-19/Australia/QLD1014/2020|EPI_ISL_530274|2020 ···················· ···················· ···················· ···················· ···················· ····················
H 4894 hCoV-19/Australia/QLD1015/2020|EPI_ISL_530275|2020 ···················· ···················· ···················· ···················· ···················· ····················
H 4895 hCoV-19/Australia/QLD1016/2020|EPI_ISL_530276|2020 ···················· ···················· ···················· ···················· ···················· ····················
H 4896 hCoV-19/Australia/QLD1017/2020|EPI_ISL_530277|2020 ···················· ···················· ···················· ···················· ···················· ····················
H 4897 hCoV-19/Australia/QLD1024/2020|EPI_ISL_530278|2020 ···················· ···················· ···················· ···················· ···················· ····················
H 4898 hCoV-19/Australia/QLD1025/2020|EPI_ISL_530279|2020 ···················· ···················· ···················· ···················· ···················· ····················
H 4899 hCoV-19/Australia/QLD1027/2020|EPI_ISL_530280|2020 ···················· ···················· ···················· ···················· ···················· ····················
H 4900 hCoV-19/Australia/QLD1029/2020|EPI_ISL_530281|2020 ···················· ···················· ···················· ···················· ···················· ····················
H 4901 hCoV-19/Australia/QLD1030/2020|EPI_ISL_530282|2020 ···················· ···················· ···················· ···················· ···················· ····················
H 4902 hCoV-19/Australia/QLD1031/2020|EPI_ISL_530283|2020 ···················· ···················· ···················· ···················· ···················· ····················
H 4903 hCoV-19/Scotland/QEUH-9633CD/2020|EPI_ISL_531577|2 ···················· ···················· ···················· ···················· ···················· ····················
H 4904 hCoV-19/England/QEUH-96A636/2020|EPI_ISL_531578|20 ···················· ···················· ···················· ···················· ···················· ····················
H 4905 hCoV-19/England/QEUH-963521/2020|EPI_ISL_531580|20 ···················· ·A·················· ···················· ···················· ···················· ····················
H 4906 hCoV-19/Northern Ireland/QEUH-9635B8/2020|EPI_ISL_ ···················· ···················· ···················· ···················· ···················· ····················
H 4907 hCoV-19/England/QEUH-962F60/2020|EPI_ISL_531582|20 ···················· ···················· ···················· ···················· ···················· ····················
H 4908 hCoV-19/Scotland/QEUH-96305D/2020|EPI_ISL_531583|2 ···················· ···················· ···················· ···················· ···················· ····················
H 4909 hCoV-19/England/QEUH-962E63/2020|EPI_ISL_531584|20 ···················· ·A·················· ···················· ···················· ···················· ··A·················
H 4910 hCoV-19/England/QEUH-9638FB/2020|EPI_ISL_531586|20 ···················· ···················· ···················· ···················· ···················· ····················
H 4911 hCoV-19/Northern Ireland/QEUH-9621A6/2020|EPI_ISL_ ···················· ···················· ···················· ···················· ···················· ····················
H 4912 hCoV-19/Northern Ireland/QEUH-9622EF/2020|EPI_ISL_ ···················· ···················· ···················· ···················· ···················· ····················
H 4913 hCoV-19/Scotland/QEUH-9629BD/2020|EPI_ISL_531590|2 ···················· ···················· ···················· ···················· ···················· ····················
H 4914 hCoV-19/England/QEUH-9632C0/2020|EPI_ISL_531592|20 ···················· ···················· ···················· ···················· ···················· ····················
H 4915 hCoV-19/England/QEUH-962ECD/2020|EPI_ISL_531593|20 ···················· ···················· ···················· ···················· ···················· ····················
H 4916 hCoV-19/Northern Ireland/QEUH-96223A/2020|EPI_ISL_ ···················· ···················· ···················· ···················· ···················· ····················
H 4917 hCoV-19/Scotland/QEUH-961FF8/2020|EPI_ISL_531595|2 ···················· ···················· ···················· ···················· ···················· ····················
H 4918 hCoV-19/England/QEUH-963758/2020|EPI_ISL_531596|20 ···················· ···················· ···················· ···················· ···················· ····················
H 4919 hCoV-19/Northern Ireland/QEUH-9622B2/2020|EPI_ISL_ ···················· ···················· ···················· ···················· ···················· ····················
H 4920 hCoV-19/Scotland/QEUH-963907/2020|EPI_ISL_531598|2 ···················· ···················· ···················· ···················· ···················· ····················
H 4921 hCoV-19/Scotland/QEUH-961FAD/2020|EPI_ISL_531599|2 ···················· ···················· ···················· ···················· ···················· ····················
H 4922 hCoV-19/Northern Ireland/QEUH-962373/2020|EPI_ISL_ ···················· ···················· ···················· ···················· ···················· ····················
H 4923 hCoV-19/England/QEUH-962E09/2020|EPI_ISL_531701|20 ···················· ···················· ···················· ···················· ···················· ····················
H 4924 hCoV-19/Scotland/QEUH-962003/2020|EPI_ISL_531702|2 ···················· ···················· ···················· ···················· ···················· ····················
H 4925 hCoV-19/Scotland/QEUH-962C78/2020|EPI_ISL_531703|2 ···················· ···················· ···················· ···················· ···················· ····················
H 4926 hCoV-19/England/QEUH-96259B/2020|EPI_ISL_531704|20 ···················· ···················· ···················· ···················· ···················· ····················
H 4927 hCoV-19/England/QEUH-962F7F/2020|EPI_ISL_531705|20 ···················· ···················· ···················· ···················· ···················· ····················
H 4928 hCoV-19/England/QEUH-962DC0/2020|EPI_ISL_531707|20 ···················· ·A·················· ···················· ···················· ···················· ····················
H 4929 hCoV-19/England/QEUH-962E81/2020|EPI_ISL_531708|20 ···················· ···················· ···················· ···················· ···················· ····················
H 4930 hCoV-19/England/QEUH-962917/2020|EPI_ISL_531709|20 ···················· ···················· ···················· ···················· ···················· ····················
H 4931 hCoV-19/England/QEUH-9635C7/2020|EPI_ISL_531710|20 ···················· ···················· ···················· ···················· ···················· ····················
H 4932 hCoV-19/Northern Ireland/QEUH-96222B/2020|EPI_ISL_ ···················· ···················· ···················· ···················· ···················· ····················
H 4933 hCoV-19/Northern Ireland/QEUH-96221C/2020|EPI_ISL_ ···················· ···················· ···················· ···················· ···················· ····················
H 4934 hCoV-19/England/QEUH-962DDF/2020|EPI_ISL_531713|20 ···················· ···················· ···················· ···················· ···················· ····················
H 4935 hCoV-19/Scotland/QEUH-9632DF/2020|EPI_ISL_531714|2 ···················· ···················· ···················· ···················· ···················· ····················
H 4936 hCoV-19/England/QEUH-9624F8/2020|EPI_ISL_531715|20 ···················· ···················· ···················· ···················· ···················· ····················
H 4937 hCoV-19/England/QEUH-962944/2020|EPI_ISL_531716|20 ···················· ···················· ···················· ···················· ···················· ····················
H 4938 hCoV-19/England/QEUH-962522/2020|EPI_ISL_531718|20 ···················· ···················· ···················· ···················· ···················· ····················
H 4939 hCoV-19/England/QEUH-96274A/2020|EPI_ISL_531719|20 ···················· ···················· ···················· ···················· ···················· ····················
H 4940 hCoV-19/England/QEUH-963363/2020|EPI_ISL_531720|20 ···················· ·A·················· ···················· ···················· ···················· ····················
H 4941 hCoV-19/England/QEUH-962AE7/2020|EPI_ISL_531722|20 ···················· ···················· ···················· ···················· ···················· ····················
H 4942 hCoV-19/Northern Ireland/QEUH-96206D/2020|EPI_ISL_ ···················· ···················· ···················· ···················· ···················· ····················
H 4943 hCoV-19/England/QEUH-9624E9/2020|EPI_ISL_531724|20 ···················· ···················· ···················· ···················· ···················· ····················
H 4944 hCoV-19/England/QEUH-962CC3/2020|EPI_ISL_531725|20 ···················· ···················· ···················· ···················· ···················· ····················
H 4945 hCoV-19/Northern Ireland/QEUH-962F15/2020|EPI_ISL_ ···················· ···················· ···················· ···················· ···················· ····················
H 4946 hCoV-19/Scotland/QEUH-962B11/2020|EPI_ISL_531730|2 ···················· ···················· ···················· ···················· ···················· ····················
H 4947 hCoV-19/England/SHEF-D281E/2020|EPI_ISL_535100|202 ···················· ···················· ···················· ···················· ···················· ····················
H 4948 hCoV-19/Scotland/CVR4002/2020|EPI_ISL_535102|2020- ···················· ···················· ···················· ···················· ···················· ····················
H 4949 hCoV-19/Scotland/CVR4003/2020|EPI_ISL_535103|2020- ···················· ···················· ···················· ···················· ···················· ····················
H 4950 hCoV-19/Scotland/CVR4005/2020|EPI_ISL_535105|2020- ···················· ···················· ···················· ···················· ···················· ····················
H 4951 hCoV-19/Scotland/CVR4006/2020|EPI_ISL_535106|2020- ···················· ···················· ···················· ···················· ···················· ····················
H 4952 hCoV-19/Scotland/CVR4007/2020|EPI_ISL_535107|2020- ···················· ···················· ···················· ···················· ···················· ····················
H 4953 hCoV-19/Scotland/CVR4008/2020|EPI_ISL_535108|2020- ···················· ···················· ···················· ···················· ···················· ····················
H 4954 hCoV-19/Scotland/CVR4010/2020|EPI_ISL_535110|2020- ···················· ·A·················· ···················· ···················· ···················· ····················
H 4955 hCoV-19/Scotland/CVR4012/2020|EPI_ISL_535111|2020- ···················· ···················· ···················· ···················· ···················· ····················
H 4956 hCoV-19/Scotland/CVR4013/2020|EPI_ISL_535112|2020- ···················· ···················· ···················· ···················· ···················· ····················
H 4957 hCoV-19/Scotland/CVR4014/2020|EPI_ISL_535113|2020- ···················· ···················· ···················· ···················· ···················· ····················
H 4958 hCoV-19/Scotland/CVR4015/2020|EPI_ISL_535114|2020- ···················· ···················· ···················· ···················· ···················· ····················
H 4959 hCoV-19/Scotland/CVR4017/2020|EPI_ISL_535116|2020- ···················· ···················· ···················· ···················· ···················· ····················
H 4960 hCoV-19/Scotland/CVR4018/2020|EPI_ISL_535117|2020- ···················· ···················· ···················· ···················· ···················· ····················
H 4961 hCoV-19/Scotland/CVR4019/2020|EPI_ISL_535118|2020- ···················· ···················· ···················· ···················· ···················· ····················
H 4962 hCoV-19/Scotland/CVR4020/2020|EPI_ISL_535119|2020- ···················· ···················· ···················· ···················· ···················· ····················
H 4963 hCoV-19/Scotland/CVR4023/2020|EPI_ISL_535122|2020- ···················· ···················· ···················· ···················· ···················· ····················
H 4964 hCoV-19/Scotland/CVR4025/2020|EPI_ISL_535124|2020- ···················· ···················· ···················· ···················· ···················· ····················
H 4965 hCoV-19/Scotland/CVR4028/2020|EPI_ISL_535127|2020- ···················· ···················· ···················· ···················· ···················· ····················
H 4966 hCoV-19/Scotland/CVR4030/2020|EPI_ISL_535128|2020- ···················· ···················· ···················· ···················· ···················· ····················
H 4967 hCoV-19/Scotland/CVR4031/2020|EPI_ISL_535129|2020- ···················· ···················· ···················· ···················· ···················· ····················
H 4968 hCoV-19/Scotland/CVR4032/2020|EPI_ISL_535130|2020- ···················· ···················· ···················· ···················· ···················· ····················
H 4969 hCoV-19/Scotland/CVR4034/2020|EPI_ISL_535132|2020- ···················· ···················· ···················· ···················· ···················· ····················
H 4970 hCoV-19/Scotland/CVR4037/2020|EPI_ISL_535134|2020- ···················· ···················· ···················· ···················· ···················· ····················

H 4971 hCoV-19/Scotland/CVR4038/2020|EPI_ISL_535135|2020- ···················· ···················· ···················· ···················· ···················· ····················
H 4972 hCoV-19/Scotland/CVR4039/2020|EPI_ISL_535136|2020- ···················· ···················· ···················· ···················· ···················· ····················
H 4973 hCoV-19/Scotland/CVR4040/2020|EPI_ISL_535137|2020- ···················· ···················· ···················· ···················· ···················· ····················
H 4974 hCoV-19/Scotland/CVR4041/2020|EPI_ISL_535138|2020- ···················· ···················· ···················· ···················· ···················· ····················
H 4975 hCoV-19/USA/NM-NMDOH-2020135001/2020|EPI_ISL_53528 ···················· ···················· ···················· ···················· ···················· ····················
H 4976 hCoV-19/USA/NM-NMDOH-2020135004/2020|EPI_ISL_53529 ···················· ···················· ···················· ···················· ···················· ····················
H 4977 hCoV-19/USA/NM-NMDOH-2020135005/2020|EPI_ISL_53529 ···················· ···················· ···················· ···················· ···················· ····················
H 4978 hCoV-19/USA/NM-NMDOH-2020135008/2020|EPI_ISL_53529 ···················· ···················· ···················· ···················· ···················· ····················
H 4979 hCoV-19/USA/NM-NMDOH-2020135016/2020|EPI_ISL_53529 ···················· ···················· ···················· ···················· ···················· ····················
H 4980 hCoV-19/USA/NM-NMDOH-2020229805/2020|EPI_ISL_53529 ···················· ···················· ···················· ···················· ···················· ····················
H 4981 hCoV-19/USA/NM-NMDOH-2020229988/2020|EPI_ISL_53529 ···················· ···················· ···················· ···················· ···················· ····················
H 4982 hCoV-19/USA/NM-NMDOH-2020229996/2020|EPI_ISL_53529 ···················· ···················· ···················· ···················· ···················· ····················
H 4983 hCoV-19/Hong Kong/MV2007-00689/2020|EPI_ISL_536411 ···················· ···················· ···················· ···················· ···················· ····················
H 4984 hCoV-19/Singapore/871/2020|EPI_ISL_536412|2020-04- ···················· ···················· ···················· ···················· ···················· ····················
H 4985 hCoV-19/Singapore/872/2020|EPI_ISL_536413|2020-05- ···················· ···················· ···················· ···················· ···················· ····················
H 4986 hCoV-19/Singapore/873/2020|EPI_ISL_536414|2020-04- ···················· ···················· ···················· ···················· ···················· ····················
H 4987 hCoV-19/Singapore/874/2020|EPI_ISL_536415|2020-04- ···················· ···················· ···················· ···················· ···················· ····················
H 4988 hCoV-19/Singapore/875/2020|EPI_ISL_536416|2020-04- ···················· ···················· ···················· ···················· ···················· ····················
H 4989 hCoV-19/Singapore/876/2020|EPI_ISL_536417|2020-08- ···················· ···················· ···················· ···················· ···················· ····················
H 4990 hCoV-19/Singapore/877/2020|EPI_ISL_536418|2020-08- ···················· ···················· ···················· ···················· ···················· ····················
H 4991 hCoV-19/Singapore/878/2020|EPI_ISL_536419|2020-08- ···················· ·A·················· ···················· ···················· ···················· ··A·················
H 4992 hCoV-19/Singapore/879/2020|EPI_ISL_536420|2020-08- ···················· ···················· ···················· ···················· ···················· ····················
H 4993 hCoV-19/Singapore/880/2020|EPI_ISL_536421|2020-08- ···················· ···················· ···················· ···················· ···················· ····················
H 4994 hCoV-19/Singapore/881/2020|EPI_ISL_536422|2020-04- ···················· ···················· ···················· ···················· ···················· ····················
H 4995 hCoV-19/Singapore/882/2020|EPI_ISL_536423|2020-08- ···················· ···················· ···················· ···················· ···················· ····················
H 4996 hCoV-19/Singapore/883/2020|EPI_ISL_536424|2020-08- ···················· ···················· ···················· ···················· ···················· ····················
H 4997 hCoV-19/Singapore/884/2020|EPI_ISL_536425|2020-08- ···················· ···················· ···················· ···················· ···················· ····················
H 4998 hCoV-19/Singapore/885/2020|EPI_ISL_536426|2020-05- ···················· ···················· ···················· ···················· ···················· ····················
H 4999 hCoV-19/Singapore/886/2020|EPI_ISL_536427|2020-08- ···················· ···················· ···················· ···················· ···················· ····················
H 5000 hCoV-19/Singapore/887/2020|EPI_ISL_536428|2020-08- ···················· ···················· ···················· ···················· ···················· ····················
H 5001 hCoV-19/Singapore/888/2020|EPI_ISL_536429|2020-09- ···················· ·A·················· ···················· ···················· ···················· ··A·················
H 5002 hCoV-19/Singapore/889/2020|EPI_ISL_536430|2020-09- ···················· ···················· ···················· ···················· ···················· ····················
H 5003 hCoV-19/Singapore/890/2020|EPI_ISL_536431|2020-09- ···················· ···················· ···················· ···················· ···················· ····················
H 5004 hCoV-19/Singapore/891/2020|EPI_ISL_536432|2020-09- ···················· ···················· ···················· ···················· ···················· ····················
H 5005 hCoV-19/Singapore/892/2020|EPI_ISL_536433|2020-09- ···················· ···················· ···················· ···················· ···················· ····················
H 5006 hCoV-19/Singapore/893/2020|EPI_ISL_536434|2020-09- ···················· ·A·················· ···················· ···················· ···················· ··A·················
H 5007 hCoV-19/Singapore/894/2020|EPI_ISL_536435|2020-02- ···················· ···················· ···················· ···················· ···················· ····················
H 5008 hCoV-19/Singapore/895/2020|EPI_ISL_536436|2020-03- ···················· ···················· ···················· ···················· ···················· ····················
H 5009 hCoV-19/Singapore/896/2020|EPI_ISL_536437|2020-03- ···················· ···················· ···················· ···················· ···················· ····················
H 5010 hCoV-19/Singapore/897/2020|EPI_ISL_536438|2020-09- ···················· ···················· ···················· ···················· ···················· ····················
H 5011 hCoV-19/Singapore/898/2020|EPI_ISL_536439|2020-02- ···················· ···················· ···················· ···················· ···················· ····················
H 5012 hCoV-19/Singapore/899/2020|EPI_ISL_536440|2020-03- ···················· ···················· ···················· ···················· ···················· ····················
H 5013 hCoV-19/Singapore/900/2020|EPI_ISL_536441|2020-02- ···················· ···················· ···················· ···················· ···················· ····················
H 5014 hCoV-19/Singapore/901/2020|EPI_ISL_536442|2020-09- ···················· ·A·················· ···················· ···················· ···················· ··A·················
H 5015 hCoV-19/Singapore/902/2020|EPI_ISL_536443|2020-08- ···················· ···················· ···················· ···················· ···················· ····················
H 5016 hCoV-19/Singapore/903/2020|EPI_ISL_536444|2020-09- ···················· ···················· ···················· ···················· ···················· ····················
H 5017 hCoV-19/Singapore/904/2020|EPI_ISL_536445|2020-09- ···················· ···················· ···················· ···················· ···················· ····················
H 5018 hCoV-19/Singapore/905/2020|EPI_ISL_536446|2020-09- ···················· ···················· ···················· ···················· ···················· ····················
H 5019 hCoV-19/Singapore/906/2020|EPI_ISL_536447|2020-01- ···················· ···················· ···················· ···················· ···················· ····················
H 5020 hCoV-19/Singapore/907/2020|EPI_ISL_536448|2020-04- ···················· ···················· ···················· ···················· ···················· ····················
H 5021 hCoV-19/Singapore/908/2020|EPI_ISL_536449|2020-09- ···················· ···················· ···················· ···················· ···················· ····················
H 5022 hCoV-19/Singapore/909/2020|EPI_ISL_536450|2020-04- ···················· ···················· ···················· ···················· ···················· ····················
H 5024 hCoV-19/Spain/CN-IBV-97006508/2020|EPI_ISL_537716| ···················· ···················· ···················· ···················· ···················· ····················
H 5027 hCoV-19/Spain/CN-IBV-97006516/2020|EPI_ISL_537721| ···················· ·A·················· ···················· ···················· ···················· ····················
H 5028 hCoV-19/Spain/CN-IBV-97006519/2020|EPI_ISL_537724| ···················· ···················· ···················· ···················· ···················· ····················
H 5029 hCoV-19/Italy/LOM-UniMI-L253/2020|EPI_ISL_542225|2 ···················· ···················· ···················· ···················· ···················· ····················
H 5030 hCoV-19/Italy/LOM-UniMI-L254/2020|EPI_ISL_542226|2 ···················· ···················· ···················· ···················· ···················· ····················
H 5031 hCoV-19/Italy/LOM-UniMI-L257/2020|EPI_ISL_542228|2 ···················· ···················· ···················· ···················· ···················· ····················
H 5032 hCoV-19/Italy/LOM-UniMI-L259/2020|EPI_ISL_542229|2 ···················· ···················· ···················· ···················· ···················· ····················
H 5033 hCoV-19/Italy/LOM-UniMI-L260/2020|EPI_ISL_542230|2 ···················· ···················· ···················· ···················· ···················· ····················
H 5034 hCoV-19/Italy/LOM-UniMI-L261/2020|EPI_ISL_542231|2 ···················· ···················· ···················· ···················· ···················· ····················
H 5035 hCoV-19/Italy/LOM-UniMI-L265/2020|EPI_ISL_542232|2 ···················· ···················· ···················· ···················· ···················· ····················
H 5036 hCoV-19/Italy/LOM-UniMI-L267/2020|EPI_ISL_542233|2 ···················· ···················· ···················· ···················· ···················· ····················
H 5037 hCoV-19/Italy/LOM-UniMI-L268/2020|EPI_ISL_542234|2 ···················· ···················· ···················· ···················· ···················· ····················
H 5038 hCoV-19/Italy/LOM-UniMI-L269/2020|EPI_ISL_542235|2 ···················· ···················· ···················· ···················· ···················· ····················
H 5039 hCoV-19/Italy/LOM-UniMI-L270/2020|EPI_ISL_542236|2 ···················· ···················· ···················· ···················· ···················· ····················
H 5040 hCoV-19/Italy/LOM-UniMI-L271/2020|EPI_ISL_542237|2 ···················· ···················· ···················· ···················· ···················· ····················
H 5041 hCoV-19/Italy/LOM-UniMI-L272/2020|EPI_ISL_542238|2 ···················· ···················· ···················· ···················· ···················· ····················
H 5042 hCoV-19/Italy/LOM-UniMI-L273/2020|EPI_ISL_542239|2 ···················· ···················· ···················· ···················· ···················· ····················
H 5043 hCoV-19/Italy/LOM-UniMI-L274/2020|EPI_ISL_542240|2 ···················· ···················· ···················· ···················· ···················· ····················
H 5044 hCoV-19/Italy/LOM-UniMI-L276/2020|EPI_ISL_542241|2 ···················· ···················· ···················· ···················· ···················· ····················
H 5045 hCoV-19/Italy/LOM-UniMI-L278/2020|EPI_ISL_542242|2 ···················· ···················· ···················· ···················· ···················· ····················
H 5046 hCoV-19/Italy/LOM-UniMI-L279/2020|EPI_ISL_542243|2 ···················· ···················· ···················· ···················· ···················· ····················
H 5047 hCoV-19/Italy/LOM-UniMI-L280/2020|EPI_ISL_542244|2 ···················· ···················· ···················· ···················· ···················· ····················
H 5048 hCoV-19/Italy/LOM-UniMI-L281/2020|EPI_ISL_542245|2 ···················· ···················· ···················· ···················· ···················· ····················
H 5049 hCoV-19/Italy/LOM-UniMI-L282/2020|EPI_ISL_542246|2 ···················· ···················· ···················· ···················· ···················· ····················
H 5050 hCoV-19/Italy/LOM-UniMI-L283/2020|EPI_ISL_542247|2 ···················· ···················· ···················· ···················· ···················· ····················
H 5051 hCoV-19/Italy/LOM-UniMI-L284/2020|EPI_ISL_542248|2 ···················· ···················· ···················· ···················· ···················· ····················
H 5052 hCoV-19/Italy/LOM-UniMI-L285/2020|EPI_ISL_542249|2 ···················· ···················· ···················· ···················· ···················· ····················
H 5053 hCoV-19/Italy/LOM-UniMI-L286/2020|EPI_ISL_542250|2 ···················· ···················· ···················· ···················· ···················· ····················

| | | | | | | | | |
|---|---|---|---|---|---|---|---|---|
| H | 5054 hCoV-19/Italy/LOM-UniMI-L287/2020\|EPI_ISL_542251\|2 | ···················· | ···················· | ···················· | ···················· | ···················· | ···················· | |
| H | 5055 hCoV-19/Italy/LOM-UniMI-L288/2020\|EPI_ISL_542252\|2 | ···················· | ···················· | ···················· | ···················· | ···················· | ···················· | |
| H | 5056 hCoV-19/Italy/LOM-UniMI-L289/2020\|EPI_ISL_542253\|2 | ···················· | ···················· | ···················· | ···················· | ···················· | ···················· | |
| H | 5057 hCoV-19/Italy/LOM-UniMI-L290/2020\|EPI_ISL_542254\|2 | ···················· | ···················· | ···················· | ···················· | ···················· | ···················· | |
| H | 5058 hCoV-19/Italy/LOM-UniMI-L292/2020\|EPI_ISL_542255\|2 | ···················· | ···················· | ···················· | ···················· | ···················· | ···················· | |
| H | 5059 hCoV-19/Italy/LOM-UniMI-L293/2020\|EPI_ISL_542256\|2 | ···················· | ···················· | ···················· | ···················· | ···················· | ···················· | |
| H | 5060 hCoV-19/Italy/LOM-UniMI-L294/2020\|EPI_ISL_542257\|2 | ···················· | ···················· | ···················· | ···················· | ···················· | ···················· | |
| H | 5061 hCoV-19/Italy/LOM-UniMI-L295/2020\|EPI_ISL_542258\|2 | ···················· | ···················· | ···················· | ···················· | ···················· | ···················· | |
| H | 5062 hCoV-19/Italy/LOM-UniMI-L296/2020\|EPI_ISL_542259\|2 | ···················· | ···················· | ···················· | ···················· | ···················· | ···················· | |
| H | 5063 hCoV-19/Italy/LOM-UniMI-L299/2020\|EPI_ISL_542260\|2 | ···················· | ···················· | ···················· | ···················· | ···················· | ···················· | |
| H | 5064 hCoV-19/Italy/LOM-UniMI-L302/2020\|EPI_ISL_542261\|2 | ···················· | ···················· | ···················· | ···················· | ···················· | ···················· | |
| H | 5065 hCoV-19/Italy/LOM-UniMI-L309/2020\|EPI_ISL_542262\|2 | ···················· | ···················· | ···················· | ···················· | ···················· | ···················· | |
| H | 5066 hCoV-19/Italy/LOM-UniMI-L310/2020\|EPI_ISL_542263\|2 | ···················· | ···················· | ···················· | ···················· | ···················· | ···················· | |
| H | 5067 hCoV-19/Italy/LOM-UniMI-L311/2020\|EPI_ISL_542264\|2 | ···················· | ···················· | ···················· | ···················· | ···················· | ···················· | |
| H | 5068 hCoV-19/Italy/LOM-UniMI-L313/2020\|EPI_ISL_542265\|2 | ···················· | ···················· | ···················· | ···················· | ···················· | ···················· | |
| H | 5069 hCoV-19/Italy/LOM-UniMI-L314/2020\|EPI_ISL_542266\|2 | ···················· | ···················· | ···················· | ···················· | ···················· | ···················· | |
| H | 5070 hCoV-19/Italy/LOM-UniMI-L316/2020\|EPI_ISL_542267\|2 | ···················· | ···················· | ···················· | ···················· | ···················· | ···················· | |
| H | 5071 hCoV-19/Italy/LOM-UniMI-L317/2020\|EPI_ISL_542268\|2 | ···················· | ···················· | ···················· | ···················· | ···················· | ···················· | |
| H | 5072 hCoV-19/Italy/LOM-UniMI-L318/2020\|EPI_ISL_542269\|2 | ···················· | ···················· | ···················· | ···················· | ···················· | ···················· | |
| H | 5073 hCoV-19/Italy/LOM-UniMI-L331/2020\|EPI_ISL_542278\|2 | ···················· | ···················· | ···················· | ···················· | ···················· | ···················· | |
| H | 5074 hCoV-19/Italy/LOM-UniMI-L334/2020\|EPI_ISL_542279\|2 | ···················· | ···················· | ···················· | ···················· | ···················· | ···················· | |
| H | 5077 hCoV-19/USA/TX-HMH-4359/2020\|EPI_ISL_543558\|2020-0 | ···················· | ···················· | ···················· | ···················· | ···················· | ···················· | |
| H | 5079 hCoV-19/USA/TX-HMH-4368/2020\|EPI_ISL_543565\|2020-0 | ···················· | ···················· | ···················· | ···················· | ···················· | ···················· | |
| H | 5081 hCoV-19/USA/TX-HMH-4370/2020\|EPI_ISL_543567\|2020-0 | ···················· | ···················· | ···················· | ···················· | ···················· | ···················· | |
| H | 5082 hCoV-19/USA/TX-HMH-4371/2020\|EPI_ISL_543568\|2020-0 | ···················· | ···················· | ···················· | ···················· | ···················· | ···················· | |
| H | 5083 hCoV-19/USA/TX-HMH-4372/2020\|EPI_ISL_543569\|2020-0 | ···················· | ···················· | ···················· | ···················· | ···················· | ···················· | |
| H | 5085 hCoV-19/USA/TX-HMH-4374/2020\|EPI_ISL_543571\|2020-0 | ···················· | ···················· | ···················· | ···················· | ···················· | ···················· | |
| H | 5086 hCoV-19/USA/TX-HMH-4375/2020\|EPI_ISL_543572\|2020-0 | ···················· | ···················· | ···················· | ···················· | ···················· | ···················· | |
| H | 5087 hCoV-19/USA/TX-HMH-4377/2020\|EPI_ISL_543574\|2020-0 | ···················· | ···················· | ···················· | ···················· | ···················· | ···················· | |
| H | 5088 hCoV-19/USA/TX-HMH-4378/2020\|EPI_ISL_543575\|2020-0 | ···················· | ···················· | ···················· | ···················· | ···················· | ···················· | |
| H | 5089 hCoV-19/USA/TX-HMH-4379/2020\|EPI_ISL_543576\|2020-0 | ···················· | ···················· | ···················· | ···················· | ···················· | ···················· | |
| H | 5092 hCoV-19/USA/TX-HMH-4386/2020\|EPI_ISL_543580\|2020-0 | ···················· | ···················· | ···················· | ···················· | ···················· | NN·N················ | 1 |
| H | 5093 hCoV-19/USA/TX-HMH-4388/2020\|EPI_ISL_543581\|2020-0 | ···················· | ···················· | ···················· | ···················· | ··············N····· | NN·················· | |
| H | 5094 hCoV-19/USA/TX-HMH-4389/2020\|EPI_ISL_543582\|2020-0 | ···················· | ···················· | ···················· | ···················· | ···················· | ···················· | |
| H | 5095 hCoV-19/USA/TX-HMH-4390/2020\|EPI_ISL_543583\|2020-0 | ···················· | ···················· | ···················· | ···················· | ···················· | ···················· | |
| H | 5097 hCoV-19/USA/TX-HMH-4395/2020\|EPI_ISL_543585\|2020-0 | ···················· | ···················· | ···················· | ···················· | ···················· | ···················· | |
| H | 5098 hCoV-19/USA/TX-HMH-4397/2020\|EPI_ISL_543586\|2020-0 | ···················· | ···················· | ···················· | ···················· | ···················· | ···················· | |
| H | 5101 hCoV-19/USA/TX-HMH-4400/2020\|EPI_ISL_543589\|2020-0 | ···················· | ···················· | ···················· | ···················· | ···················· | ···················· | |
| H | 5102 hCoV-19/USA/TX-HMH-4401/2020\|EPI_ISL_543590\|2020-0 | ···················· | ···················· | ···················· | ···················· | ···················· | ···················· | |
| H | 5103 hCoV-19/USA/TX-HMH-4402/2020\|EPI_ISL_543591\|2020-0 | ···················· | ···················· | ···················· | ···················· | ···················· | ···················· | |
| H | 5106 hCoV-19/USA/TX-HMH-4405/2020\|EPI_ISL_543594\|2020-0 | ···················· | ···················· | ···················· | ···················· | ···················· | ···················· | |
| H | 5108 hCoV-19/USA/TX-HMH-4408/2020\|EPI_ISL_543597\|2020-0 | ···················· | ···················· | ···················· | ···················· | ···················· | ···················· | |
| H | 5109 hCoV-19/USA/TX-HMH-4409/2020\|EPI_ISL_543598\|2020-0 | ···················· | ···················· | ···················· | ···················· | ···················· | ···················· | |
| H | 5110 hCoV-19/USA/TX-HMH-4410/2020\|EPI_ISL_543599\|2020-0 | ···················· | ·············A······ | ···················· | ···················· | ···················· | N··················· | |
| H | 5111 hCoV-19/USA/TX-HMH-4534/2020\|EPI_ISL_543700\|2020-0 | ···················· | ···················· | ···················· | ···················· | ···················· | ···················· | |
| H | 5112 hCoV-19/USA/TX-HMH-4536/2020\|EPI_ISL_543701\|2020-0 | ···················· | ···················· | ···················· | ···················· | ···················· | ·······N············ | |
| H | 5114 hCoV-19/USA/TX-HMH-454/2020\|EPI_ISL_543703\|2020-04 | ···················· | ···················· | ···················· | ···················· | ···················· | ···················· | |
| H | 5115 hCoV-19/USA/TX-HMH-4540/2020\|EPI_ISL_543704\|2020-0 | ···················· | ···················· | ···················· | ···················· | ··············N····· | NN·N················ | 1 |
| H | 5117 hCoV-19/USA/TX-HMH-4542/2020\|EPI_ISL_543706\|2020-0 | ···················· | ···················· | ···················· | ···················· | ···················· | ···················· | |
| H | 5120 hCoV-19/USA/TX-HMH-4546/2020\|EPI_ISL_543709\|2020-0 | ···················· | ···················· | ···················· | ···················· | ··············N····· | ···················· | |
| H | 5121 hCoV-19/Australia/NSW2722/2020\|EPI_ISL_530334\|2020 | ···················· | ···················· | ···················· | ···················· | ···················· | ···················· | |
| H | 5122 hCoV-19/Australia/NSW2723/2020\|EPI_ISL_530335\|2020 | ···················· | ···················· | ···················· | ···················· | ···················· | ···················· | |
| H | 5123 hCoV-19/Australia/NSW2738/2020\|EPI_ISL_530336\|2020 | ···················· | ···················· | ···················· | ···················· | ···················· | ···················· | |
| H | 5124 hCoV-19/Australia/NSW2739/2020\|EPI_ISL_530337\|2020 | ···················· | ···················· | ···················· | ···················· | ···················· | ···················· | |
| H | 5125 hCoV-19/Australia/NSW2741/2020\|EPI_ISL_530338\|2020 | ···················· | ···················· | ···················· | ···················· | ···················· | ···················· | |
| H | 5126 hCoV-19/Australia/NSW2749/2020\|EPI_ISL_530339\|2020 | ···················· | ···················· | ···················· | ···················· | ···················· | ···················· | |
| H | 5127 hCoV-19/Australia/NSW2750/2020\|EPI_ISL_530340\|2020 | ···················· | ···················· | ···················· | ···················· | ···················· | ···················· | |
| H | 5128 hCoV-19/Czech Republic/NRL_4038/2020\|EPI_ISL_53034 | ···················· | ···················· | ···················· | ···················· | ···················· | ···················· | |
| H | 5129 hCoV-19/Czech Republic/NRL_4182/2020\|EPI_ISL_53034 | ···················· | ···················· | ···················· | ···················· | ···················· | ···················· | |
| H | 5130 hCoV-19/Czech Republic/NRL_4262/2020\|EPI_ISL_53034 | ···················· | ···················· | ···················· | ···················· | ···················· | ···················· | |
| H | 5131 hCoV-19/Czech Republic/NRL_5553/2020\|EPI_ISL_53035 | ···················· | ···················· | ···················· | ···················· | ···················· | ···················· | |
| H | 5132 hCoV-19/England/QEUH-9904A6/2020\|EPI_ISL_530352\|20 | ···················· | ·A·················· | ···················· | ···················· | ···················· | ···················· | |
| H | 5133 hCoV-19/England/QEUH-990752/2020\|EPI_ISL_530353\|20 | ···················· | ···················· | ···················· | ···················· | ···················· | ···················· | |
| H | 5134 hCoV-19/England/QEUH-98FF9A/2020\|EPI_ISL_530358\|20 | ···················· | ·A·················· | ···················· | ···················· | ···················· | ···················· | |
| H | 5135 hCoV-19/England/QEUH-962D84/2020\|EPI_ISL_531610\|20 | ···················· | ·A·················· | ···················· | ···················· | ···················· | ···················· | |
| H | 5136 hCoV-19/Northern Ireland/QEUH-963248/2020\|EPI_ISL_ | ···················· | ···················· | ···················· | ···················· | ···················· | ···················· | |
| H | 5137 hCoV-19/England/QEUH-9625D7/2020\|EPI_ISL_531612\|20 | ···················· | ·A·················· | ···················· | ···················· | ···················· | ··A················· | |
| H | 5138 hCoV-19/Wales/QEUH-962FF7/2020\|EPI_ISL_531613\|2020 | ···················· | ···················· | ···················· | ···················· | ···················· | ···················· | |
| H | 5139 hCoV-19/England/QEUH-9634F7/2020\|EPI_ISL_531614\|20 | ···················· | ···················· | ···················· | ···················· | ···················· | ···················· | |
| H | 5140 hCoV-19/England/QEUH-9632A2/2020\|EPI_ISL_531616\|20 | ···················· | ···················· | ···················· | ···················· | ···················· | ···················· | |
| H | 5141 hCoV-19/Scotland/QEUH-963354/2020\|EPI_ISL_531617\|2 | ···················· | ···················· | ···················· | ···················· | ···················· | ···················· | |
| H | 5142 hCoV-19/England/QEUH-9626B6/2020\|EPI_ISL_531619\|20 | ···················· | ···················· | ···················· | ···················· | ···················· | ···················· | |
| H | 5143 hCoV-19/England/QEUH-96271D/2020\|EPI_ISL_531621\|20 | ···················· | ···················· | ···················· | ···················· | ···················· | ···················· | |
| H | 5144 hCoV-19/Northern Ireland/QEUH-963406/2020\|EPI_ISL_ | ···················· | ···················· | ···················· | ···················· | ···················· | ···················· | |
| H | 5145 hCoV-19/England/QEUH-962D0C/2020\|EPI_ISL_531623\|20 | ···················· | ···················· | ···················· | ···················· | ···················· | ···················· | |
| H | 5146 hCoV-19/Northern Ireland/QEUH-96208B/2020\|EPI_ISL_ | ···················· | ···················· | ···················· | ···················· | ···················· | ···················· | |
| H | 5147 hCoV-19/Scotland/QEUH-96280B/2020\|EPI_ISL_531625\|2 | ···················· | ···················· | ···················· | ···················· | ···················· | ···················· | |
| H | 5148 hCoV-19/Northern Ireland/QEUH-96220D/2020\|EPI_ISL_ | ···················· | ···················· | ···················· | ···················· | ···················· | ···················· | |
| H | 5149 hCoV-19/England/QEUH-96320C/2020\|EPI_ISL_531628\|20 | ···················· | ···················· | ···················· | ···················· | ···················· | ···················· | |
| H | 5150 hCoV-19/England/QEUH-9634D9/2020\|EPI_ISL_531629\|20 | ···················· | ···················· | ···················· | ···················· | ···················· | ···················· | |

H 5151 hCoV-19/Wales/QEUH-9635A9/2020|EPI_ISL_531630|2020 ···················· ···················· ···················· ···················· ···················· ····················
H 5152 hCoV-19/Scotland/QEUH-961FDA/2020|EPI_ISL_531631|2 ···················· ···················· ···················· ···················· ···················· ····················
H 5153 hCoV-19/England/QEUH-963916/2020|EPI_ISL_531632|20 ···················· ···················· ···················· ···················· ···················· ····················
H 5154 hCoV-19/England/QEUH-9633BE/2020|EPI_ISL_531633|20 ···················· ···················· ···················· ···················· ···················· ····················
H 5155 hCoV-19/Scotland/QEUH-962E45/2020|EPI_ISL_531634|2 ···················· ···················· ···················· ···················· ···················· ····················
H 5156 hCoV-19/England/QEUH-9633DC/2020|EPI_ISL_531635|20 ···················· ···················· ···················· ···················· ···················· ····················
H 5157 hCoV-19/Northern Ireland/QEUH-962980/2020|EPI_ISL_ ···················· ···················· ···················· ···················· ···················· ····················
H 5158 hCoV-19/Scotland/QEUH-96363D/2020|EPI_ISL_531638|2 ···················· ···················· ···················· ···················· ···················· ····················
H 5159 hCoV-19/Scotland/QEUH-962276/2020|EPI_ISL_531639|2 ···················· ···················· ···················· ···················· ···················· ····················
H 5160 hCoV-19/England/QEUH-9635F4/2020|EPI_ISL_531642|20 ···················· ···················· ···················· ···················· ···················· ····················
H 5161 hCoV-19/Northern Ireland/QEUH-9630B7/2020|EPI_ISL_ ···················· ···················· ···················· ···················· ···················· ····················
H 5162 hCoV-19/Northern Ireland/QEUH-962258/2020|EPI_ISL_ ···················· ···················· ···················· ···················· ···················· ····················
H 5163 hCoV-19/England/QEUH-961F52/2020|EPI_ISL_531645|20 ···················· ···················· ···················· ···················· ···················· ····················
H 5164 hCoV-19/England/QEUH-9633AF/2020|EPI_ISL_531647|20 ···················· ···················· ···················· ···················· ···················· ····················
H 5165 hCoV-19/Scotland/QEUH-962C69/2020|EPI_ISL_531649|2 ···················· ···················· ···················· ···················· ···················· ····················
H 5166 hCoV-19/England/QEUH-96358B/2020|EPI_ISL_531650|20 ···················· ···················· ···················· ···················· ···················· ····················
H 5167 hCoV-19/England/QEUH-962A50/2020|EPI_ISL_531651|20 ···················· ···················· ···················· ···················· ···················· ····················
H 5168 hCoV-19/Northern Ireland/QEUH-96209A/2020|EPI_ISL_ ···················· ···················· ···················· ···················· ···················· ····················
H 5169 hCoV-19/Northern Ireland/QEUH-961FCB/2020|EPI_ISL_ ···················· ···················· ···················· ···················· ···················· ····················
H 5170 hCoV-19/England/QEUH-9624DA/2020|EPI_ISL_531656|20 ···················· ···················· ···················· ···················· ···················· ····················
H 5171 hCoV-19/England/QEUH-96257D/2020|EPI_ISL_531658|20 ···················· ···················· ···················· ···················· ···················· ····················
H 5172 hCoV-19/Scotland/QEUH-965AE4/2020|EPI_ISL_531660|2 ···················· ···················· ···················· ···················· ···················· ····················
H 5173 hCoV-19/Northern Ireland/QEUH-962319/2020|EPI_ISL_ ···················· ···················· ···················· ···················· ···················· ····················
H 5174 hCoV-19/England/QEUH-9637B2/2020|EPI_ISL_531662|20 ···················· ···················· ···················· ···················· ···················· ····················
H 5175 hCoV-19/Northern Ireland/QEUH-962346/2020|EPI_ISL_ ···················· ···················· ···················· ···················· ···················· ····················
H 5176 hCoV-19/England/QEUH-962A23/2020|EPI_ISL_531664|20 ···················· ···················· ···················· ···················· ···················· ····················
H 5177 hCoV-19/England/QEUH-962B7B/2020|EPI_ISL_531665|20 ···················· ···················· ···················· ···················· ···················· ····················
H 5178 hCoV-19/England/QEUH-962CD2/2020|EPI_ISL_531666|20 ···················· ···················· ···················· ···················· ···················· ····················
H 5179 hCoV-19/England/QEUH-96267A/2020|EPI_ISL_531667|20 ···················· ···················· ···················· ···················· ···················· ····················
H 5180 hCoV-19/Scotland/QEUH-9631F0/2020|EPI_ISL_531668|2 ···················· ···················· ···················· ···················· ···················· ····················
H 5181 hCoV-19/England/QEUH-9629EA/2020|EPI_ISL_531669|20 ···················· ···················· ···················· ···················· ···················· ····················
H 5182 hCoV-19/England/QEUH-962AC9/2020|EPI_ISL_531671|20 ···················· ···················· ···················· ···················· ···················· ····················
H 5183 hCoV-19/Scotland/QEUH-962FCA/2020|EPI_ISL_531672|2 ···················· ···················· ···················· ···················· ···················· ····················
H 5184 hCoV-19/England/QEUH-963600/2020|EPI_ISL_531673|20 ···················· ···················· ···················· ···················· ···················· ····················
H 5185 hCoV-19/Northern Ireland/QEUH-96211F/2020|EPI_ISL_ ···················· ···················· ···················· ···················· ···················· ····················
H 5186 hCoV-19/England/QEUH-962759/2020|EPI_ISL_531676|20 ···················· ···················· ···················· ···················· ···················· ····················
H 5187 hCoV-19/Scotland/QEUH-9637EF/2020|EPI_ISL_531677|2 ···················· ···················· ···················· ···················· ···················· ····················
H 5188 hCoV-19/Scotland/QEUH-9631D2/2020|EPI_ISL_531678|2 ···················· ···················· ···················· ···················· ···················· ····················
H 5189 hCoV-19/England/QEUH-962A14/2020|EPI_ISL_531679|20 ···················· ···················· ···················· ···················· ···················· ····················
H 5190 hCoV-19/England/QEUH-9621E2/2020|EPI_ISL_531680|20 ···················· ·A·················· ···················· ···················· ···················· ··A·················
H 5191 hCoV-19/England/QEUH-96299F/2020|EPI_ISL_531682|20 ···················· ·A·················· ···················· ···················· ···················· ····················
H 5192 hCoV-19/England/QEUH-962CA5/2020|EPI_ISL_531683|20 ···················· ···················· ···················· ···················· ···················· ····················
H 5193 hCoV-19/England/QEUH-963819/2020|EPI_ISL_531684|20 ···················· ·A·················· ···················· ···················· ···················· ··A·················
H 5194 hCoV-19/England/QEUH-962DFD/2020|EPI_ISL_531685|20 ···················· ···················· ···················· ···················· ···················· ····················
H 5195 hCoV-19/Scotland/QEUH-962531/2020|EPI_ISL_531689|2 ···················· ···················· ···················· ···················· ···················· ····················
H 5196 hCoV-19/England/QEUH-7B903C/2020|EPI_ISL_532941|20 ···················· ···················· ···················· ···················· ···················· ····················
H 5197 hCoV-19/England/QEUH-7DAB61/2020|EPI_ISL_532942|20 ···················· ···················· ···················· ···················· ···················· ····················
H 5198 hCoV-19/Northern Ireland/QEUH-7FEE6E/2020|EPI_ISL_ ···················· ···················· ···················· ···················· ···················· ····················
H 5199 hCoV-19/England/QEUH-7BB805/2020|EPI_ISL_532945|20 ···················· ···················· ···················· ···················· ···················· ····················
H 5200 hCoV-19/England/QEUH-7E5CBB/2020|EPI_ISL_532947|20 ···················· ···················· ···················· ···················· ···················· ····················
H 5201 hCoV-19/Northern Ireland/QEUH-7F1B91/2020|EPI_ISL_ ···················· ···················· ···················· ···················· ···················· ····················
H 5202 hCoV-19/Northern Ireland/QEUH-7F138A/2020|EPI_ISL_ ···················· ···················· ···················· ···················· ···················· ····················
H 5203 hCoV-19/England/QEUH-7D447B/2020|EPI_ISL_532952|20 ···················· ···················· ···················· ···················· ···················· ····················
H 5204 hCoV-19/England/QEUH-7D35C4/2020|EPI_ISL_532953|20 ···················· ···················· ···················· ···················· ···················· ····················
H 5205 hCoV-19/England/QEUH-7B8F21/2020|EPI_ISL_532954|20 ···················· ···················· ···················· ···················· ···················· ····················
H 5206 hCoV-19/England/QEUH-7E54E0/2020|EPI_ISL_532955|20 ···················· ···················· ···················· ···················· ···················· ····················
H 5207 hCoV-19/England/QEUH-7B6767/2020|EPI_ISL_532956|20 ···················· ···················· ···················· ···················· ···················· ····················
H 5208 hCoV-19/England/QEUH-7F8FAD/2020|EPI_ISL_532957|20 ···················· ···················· ···················· ···················· ···················· ····················
H 5209 hCoV-19/England/QEUH-7FDAB6/2020|EPI_ISL_532958|20 ···················· ···················· ···················· ···················· ···················· ····················
H 5210 hCoV-19/England/QEUH-7C9C08/2020|EPI_ISL_532959|20 ···················· ···················· ···················· ···················· ···················· ····················
H 5211 hCoV-19/England/QEUH-7E2735/2020|EPI_ISL_532960|20 ···················· ···················· ···················· ···················· ···················· ····················
H 5212 hCoV-19/England/QEUH-7B655E/2020|EPI_ISL_532961|20 ···················· ···················· ···················· ···················· ···················· ····················
H 5213 hCoV-19/Scotland/QEUH-7BADF8/2020|EPI_ISL_532962|2 ···················· ···················· ···················· ···················· ···················· ····················
H 5214 hCoV-19/England/QEUH-7B8F9A/2020|EPI_ISL_532963|20 ···················· ···················· ···················· ···················· ···················· ····················
H 5215 hCoV-19/England/QEUH-75939F/2020|EPI_ISL_532964|20 ···················· ···················· ···················· ···················· ···················· ····················
H 5216 hCoV-19/England/QEUH-7675B6/2020|EPI_ISL_532965|20 ···················· ···················· ···················· ···················· ···················· ····················
H 5217 hCoV-19/England/QEUH-768694/2020|EPI_ISL_532966|20 ···················· ···················· ···················· ···················· ···················· ····················
H 5218 hCoV-19/England/QEUH-757908/2020|EPI_ISL_532967|20 ···················· ···················· ···················· ···················· ···················· ····················
H 5219 hCoV-19/England/QEUH-7689D7/2020|EPI_ISL_532968|20 ···················· ···················· ···················· ···················· ···················· ····················
H 5220 hCoV-19/England/QEUH-768C1A/2020|EPI_ISL_532970|20 ···················· ···················· ···················· ···················· ···················· ····················
H 5221 hCoV-19/England/QEUH-757470/2020|EPI_ISL_532971|20 ···················· ···················· ···················· ···················· ···················· ····················
H 5222 hCoV-19/England/QEUH-75FCBC/2020|EPI_ISL_532972|20 ···················· ···················· ···················· ···················· ···················· ····················
H 5223 hCoV-19/England/QEUH-761D0F/2020|EPI_ISL_532973|20 ···················· ···················· ···················· ···················· ···················· ····················
H 5224 hCoV-19/England/QEUH-7664AB/2020|EPI_ISL_532974|20 ···················· ···················· ···················· ···················· ···················· ····················
H 5225 hCoV-19/England/QEUH-760ED0/2020|EPI_ISL_532975|20 ···················· ···················· ···················· ···················· ···················· ····················
H 5226 hCoV-19/England/QEUH-760447/2020|EPI_ISL_532977|20 ···················· ·············A······ ···················· ···················· ···················· ····················
H 5227 hCoV-19/Scotland/QEUH-759739/2020|EPI_ISL_532978|2 ···················· ···················· ···················· ···················· ···················· ····················
H 5228 hCoV-19/Northern Ireland/QEUH-762B13/2020|EPI_ISL_ ··················T· ···················· ···················· ···················· ···················· ···················· 1
H 5229 hCoV-19/England/QEUH-7532A7/2020|EPI_ISL_532980|20 ···················· ···················· ···················· ···················· ···················· ····················
H 5230 hCoV-19/England/QEUH-75E998/2020|EPI_ISL_532981|20 ···················· ···················· ···················· ···················· ···················· ····················

| | | | | | | | | |
|---|---|---|---|---|---|---|---|---|
| H | 5231 | hCoV-19/Northern Ireland/QEUH-764CB4/2020\|EPI_ISL_ | ···················· | ···················· | ····················· | ···················· | ···················· | ···················· |
| H | 5232 | hCoV-19/Scotland/QEUH-759502/2020\|EPI_ISL_532983\|2 | ···················· | ···················· | ····················· | ···················· | ···················· | ···················· |
| H | 5233 | hCoV-19/England/QEUH-761D5A/2020\|EPI_ISL_532984\|20 | ···················· | ···················· | ····················· | ···················· | ···················· | ···················· |
| H | 5234 | hCoV-19/Northern Ireland/QEUH-7550AB/2020\|EPI_ISL_ | ···················· | ···················· | ····················· | ···················· | ···················· | ···················· |
| H | 5235 | hCoV-19/England/QEUH-76672A/2020\|EPI_ISL_532986\|20 | ···················· | ···················· | ····················· | ···················· | ···················· | ···················· |
| H | 5236 | hCoV-19/England/QEUH-75F40F/2020\|EPI_ISL_532987\|20 | ···················· | ···················· | ····················· | ···················· | ···················· | ···················· |
| H | 5237 | hCoV-19/Scotland/QEUH-755533/2020\|EPI_ISL_532988\|2 | ···················· | ···················· | ····················· | ···················· | ···················· | ···················· |
| H | 5238 | hCoV-19/England/QEUH-7668DC/2020\|EPI_ISL_532989\|20 | ···················· | ···················· | ····················· | ···················· | ···················· | ···················· |
| H | 5239 | hCoV-19/Scotland/QEUH-75958A/2020\|EPI_ISL_532990\|2 | ···················· | ···················· | ····················· | ···················· | ···················· | ···················· |
| H | 5240 | hCoV-19/England/QEUH-758BB6/2020\|EPI_ISL_532991\|20 | ···················· | ···················· | ····················· | ···················· | ···················· | ···················· |
| H | 5241 | hCoV-19/Scotland/QEUH-759F40/2020\|EPI_ISL_532992\|2 | ···················· | ···················· | ····················· | ···················· | ···················· | ···················· |
| H | 5242 | hCoV-19/England/QEUH-7631F2/2020\|EPI_ISL_532993\|20 | ···················· | ···················· | ····················· | ···················· | ···················· | ···················· |
| H | 5243 | hCoV-19/Scotland/QEUH-758FD8/2020\|EPI_ISL_532994\|2 | ···················· | ···················· | ····················· | ···················· | ···················· | ···················· |
| H | 5244 | hCoV-19/England/QEUH-75F0B0/2020\|EPI_ISL_532995\|20 | ···················· | ···················· | ····················· | ···················· | ··········T········· | ···················· |
| H | 5245 | hCoV-19/England/QEUH-757C78/2020\|EPI_ISL_532996\|20 | ···················· | ···················· | ····················· | ···················· | ···················· | ···················· |
| H | 5246 | hCoV-19/Northern Ireland/QEUH-768333/2020\|EPI_ISL_ | ···················· | ···················· | ····················· | ···················· | ···················· | ···················· |
| H | 5247 | hCoV-19/England/QEUH-762111/2020\|EPI_ISL_532998\|20 | ···················· | ···················· | ····················· | ···················· | ···················· | ···················· |
| H | 5248 | hCoV-19/England/QEUH-753EC1/2020\|EPI_ISL_532999\|20 | ···················· | ···················· | ····················· | ···················· | ···················· | ···················· |
| H | 5249 | hCoV-19/Spain/CN-IBV-97006485/2020\|EPI_ISL_537700\| | ···················· | ···················· | ····················· | ···················· | ···················· | ···················· |
| H | 5255 | hCoV-19/South Korea/IHC2681/2020\|EPI_ISL_541000\|20 | ···················· | ···················· | ····················· | ···················· | ···················· | ···················· |
| H | 5256 | hCoV-19/South Korea/IHC3486/2020\|EPI_ISL_541001\|20 | ···················· | ···················· | ····················· | ···················· | ···················· | ···················· |
| H | 5257 | hCoV-19/South Korea/IHC17695/2020\|EPI_ISL_541002\|2 | ···················· | ···················· | ····················· | ···················· | ···················· | ···················· |
| H | 5258 | hCoV-19/South Korea/IHC18094/2020\|EPI_ISL_541003\|2 | ···················· | ···················· | ····················· | ···················· | ···················· | ···················· |
| H | 5259 | hCoV-19/Italy/LOM-UniMI-L319/2020\|EPI_ISL_542270\|2 | ···················· | ···················· | ····················· | ···················· | ···················· | ···················· |
| H | 5260 | hCoV-19/Italy/LOM-UniMI-L321/2020\|EPI_ISL_542271\|2 | ···················· | ···················· | ····················· | ···················· | ···················· | ···················· |
| H | 5261 | hCoV-19/Italy/LOM-UniMI-L322/2020\|EPI_ISL_542272\|2 | ···················· | ···················· | ····················· | ···················· | ···················· | ···················· |
| H | 5262 | hCoV-19/Italy/LOM-UniMI-L325/2020\|EPI_ISL_542273\|2 | ···················· | ···················· | ····················· | ···················· | ···················· | ···················· |
| H | 5263 | hCoV-19/Italy/LOM-UniMI-L327/2020\|EPI_ISL_542274\|2 | ···················· | ···················· | ····················· | ···················· | ···················· | ···················· |
| H | 5264 | hCoV-19/Italy/LOM-UniMI-L328/2020\|EPI_ISL_542275\|2 | ···················· | ···················· | ····················· | ···················· | ···················· | ···················· |
| H | 5265 | hCoV-19/Italy/LOM-UniMI-L329/2020\|EPI_ISL_542276\|2 | ···················· | ···················· | ····················· | ···················· | ···················· | ···················· |
| H | 5266 | hCoV-19/Italy/LOM-UniMI-L330/2020\|EPI_ISL_542277\|2 | ···················· | ···················· | ····················· | ···················· | ···················· | ···················· |
| H | 5267 | hCoV-19/Italy/LOM-UniMI-L335/2020\|EPI_ISL_542280\|2 | ···················· | ···················· | ····················· | ···················· | ···················· | ···················· |
| H | 5268 | hCoV-19/Italy/LOM-UniMI-L342/2020\|EPI_ISL_542281\|2 | ···················· | ···················· | ····················· | ···················· | ···················· | ···················· |
| H | 5269 | hCoV-19/Italy/LOM-UniMI-L343/2020\|EPI_ISL_542282\|2 | ···················· | ···················· | ····················· | ···················· | ···················· | ···················· |
| H | 5270 | hCoV-19/Italy/LOM-UniMI-L345/2020\|EPI_ISL_542283\|2 | ···················· | ···················· | ····················· | ···················· | ···················· | ···················· |
| H | 5271 | hCoV-19/Italy/LOM-UniMI-L346/2020\|EPI_ISL_542284\|2 | ···················· | ···················· | ····················· | ···················· | ···················· | ···················· |
| H | 5272 | hCoV-19/Italy/LOM-UniMI-L347/2020\|EPI_ISL_542285\|2 | ···················· | ···················· | ····················· | ···················· | ···················· | ···················· |
| H | 5273 | hCoV-19/Italy/LOM-UniMI-L348/2020\|EPI_ISL_542286\|2 | ···················· | ···················· | ····················· | ···················· | ···················· | ···················· |
| H | 5274 | hCoV-19/Italy/LOM-UniMI-L350/2020\|EPI_ISL_542287\|2 | ···················· | ···················· | ····················· | ···················· | ···················· | ···················· |
| H | 5275 | hCoV-19/Italy/LOM-UniMI-L352/2020\|EPI_ISL_542288\|2 | ···················· | ···················· | ····················· | ···················· | ···················· | ···················· |
| H | 5276 | hCoV-19/Italy/LOM-UniMI-L354/2020\|EPI_ISL_542289\|2 | ···················· | ···················· | ····················· | ···················· | ···················· | ···················· |
| H | 5277 | hCoV-19/Italy/LOM-UniMI-L356/2020\|EPI_ISL_542290\|2 | ···················· | ···················· | ····················· | ···················· | ···················· | ···················· |
| H | 5278 | hCoV-19/Italy/LOM-UniMI-L357/2020\|EPI_ISL_542291\|2 | ···················· | ···················· | ····················· | ···················· | ···················· | ···················· |
| H | 5279 | hCoV-19/Italy/LOM-UniMI-L358/2020\|EPI_ISL_542292\|2 | ···················· | ···················· | ····················· | ···················· | ···················· | ···················· |
| H | 5280 | hCoV-19/Italy/LOM-UniMI-L361/2020\|EPI_ISL_542293\|2 | ···················· | ···················· | ····················· | ···················· | ···················· | ···················· |
| H | 5281 | hCoV-19/Italy/LOM-UniMI-L362/2020\|EPI_ISL_542294\|2 | ···················· | ···················· | ····················· | ···················· | ···················· | ···················· |
| H | 5282 | hCoV-19/Italy/LOM-UniMI-L363/2020\|EPI_ISL_542295\|2 | ···················· | ···················· | ····················· | ···················· | ···················· | ···················· |
| H | 5283 | hCoV-19/Italy/LOM-UniMI-L364/2020\|EPI_ISL_542296\|2 | ···················· | ···················· | ····················· | ···················· | ···················· | ···················· |
| H | 5284 | hCoV-19/Italy/LOM-UniMI-L366/2020\|EPI_ISL_542297\|2 | ···················· | ···················· | ····················· | ···················· | ···················· | ···················· |
| H | 5285 | hCoV-19/Italy/LOM-UniMI-L367/2020\|EPI_ISL_542298\|2 | ···················· | ···················· | ····················· | ···················· | ···················· | ···················· |
| H | 5286 | hCoV-19/Italy/LOM-UniMI-L368/2020\|EPI_ISL_542299\|2 | ···················· | ···················· | ····················· | ···················· | ···················· | ···················· |
| H | 5287 | hCoV-19/Italy/LOM-UniMI-L369/2020\|EPI_ISL_542300\|2 | ···················· | ···················· | ····················· | ···················· | ···················· | ···················· |
| H | 5288 | hCoV-19/Italy/LOM-UniMI-L370/2020\|EPI_ISL_542301\|2 | ···················· | ···················· | ····················· | ···················· | ···················· | ···················· |
| H | 5289 | hCoV-19/Italy/LOM-UniMI-L371/2020\|EPI_ISL_542302\|2 | ···················· | ···················· | ····················· | ···················· | ···················· | ···················· |
| H | 5290 | hCoV-19/Italy/LOM-UniMI-L372/2020\|EPI_ISL_542303\|2 | ···················· | ···················· | ····················· | ···················· | ···················· | ···················· |
| H | 5291 | hCoV-19/Italy/LOM-UniMI-L373/2020\|EPI_ISL_542304\|2 | ···················· | ···················· | ····················· | ···················· | ···················· | ···················· |
| H | 5292 | hCoV-19/Italy/LOM-UniMI-L378/2020\|EPI_ISL_542305\|2 | ···················· | ···················· | ····················· | ···················· | ···················· | ···················· |
| H | 5293 | hCoV-19/Italy/LOM-UniMI-L380/2020\|EPI_ISL_542306\|2 | ···················· | ···················· | ····················· | ···················· | ···················· | ···················· |
| H | 5294 | hCoV-19/Italy/LOM-UniMI-L384/2020\|EPI_ISL_542307\|2 | ···················· | ···················· | ····················· | ···················· | ···················· | ···················· |
| H | 5295 | hCoV-19/Italy/LOM-UniMI-L385/2020\|EPI_ISL_542308\|2 | ···················· | ···················· | ····················· | ···················· | ···················· | ···················· |
| H | 5296 | hCoV-19/Italy/LOM-UniMI-L386/2020\|EPI_ISL_542309\|2 | ···················· | ···················· | ····················· | ···················· | ···················· | ···················· |
| H | 5297 | hCoV-19/Italy/LOM-UniMI-L387/2020\|EPI_ISL_542310\|2 | ···················· | ···················· | ····················· | ···················· | ···················· | ···················· |
| H | 5298 | hCoV-19/Italy/LOM-UniMI-L388/2020\|EPI_ISL_542311\|2 | ···················· | ···················· | ····················· | ···················· | ···················· | ···················· |
| H | 5299 | hCoV-19/Italy/LOM-UniMI-L389/2020\|EPI_ISL_542312\|2 | ···················· | ···················· | ····················· | ···················· | ···················· | ···················· |
| H | 5300 | hCoV-19/Italy/LOM-UniMI-L390/2020\|EPI_ISL_542313\|2 | ···················· | ···················· | ····················· | ···················· | ···················· | ···················· |
| H | 5301 | hCoV-19/Italy/LOM-UniMI-L391/2020\|EPI_ISL_542314\|2 | ···················· | ···················· | ····················· | ···················· | ···················· | ···················· |
| H | 5302 | hCoV-19/Italy/LOM-UniMI-L393/2020\|EPI_ISL_542315\|2 | ···················· | ···················· | ····················· | ···················· | ···················· | ···················· |
| H | 5303 | hCoV-19/Italy/LOM-UniMI-L394/2020\|EPI_ISL_542316\|2 | ···················· | ···················· | ····················· | ···················· | ···················· | ···················· |
| H | 5304 | hCoV-19/Italy/LOM-UniMI-L395/2020\|EPI_ISL_542317\|2 | ···················· | ···················· | ····················· | ···················· | ···················· | ···················· |
| H | 5305 | hCoV-19/Italy/LOM-UniMI-L396/2020\|EPI_ISL_542318\|2 | ···················· | ···················· | ····················· | ···················· | ···················· | ···················· |
| H | 5306 | hCoV-19/Italy/LOM-UniMI-L397/2020\|EPI_ISL_542319\|2 | ···················· | ···················· | ····················· | ···················· | ···················· | ···················· |
| H | 5307 | hCoV-19/Italy/LOM-UniMI-L398/2020\|EPI_ISL_542320\|2 | ···················· | ···················· | ····················· | ···················· | ···················· | ···················· |
| H | 5308 | hCoV-19/Italy/LOM-UniMI-L399/2020\|EPI_ISL_542321\|2 | ···················· | ···················· | ····················· | ···················· | ···················· | ···················· |
| H | 5309 | hCoV-19/Italy/LOM-UniMI-L400/2020\|EPI_ISL_542322\|2 | ···················· | ···················· | ····················· | ···················· | ···················· | ···················· |
| H | 5310 | hCoV-19/Italy/LOM-UniMI-L401/2020\|EPI_ISL_542323\|2 | ···················· | ···················· | ····················· | ···················· | ···················· | ···················· |
| H | 5311 | hCoV-19/Italy/LOM-UniMI-L402/2020\|EPI_ISL_542324\|2 | ···················· | ···················· | ····················· | ···················· | ···················· | ···················· |
| H | 5312 | hCoV-19/Italy/LOM-UniMI-L403/2020\|EPI_ISL_542325\|2 | ···················· | ···················· | ····················· | ···················· | ···················· | ···················· |
| H | 5313 | hCoV-19/Italy/LOM-UniMI-L404/2020\|EPI_ISL_542326\|2 | ···················· | ···················· | ····················· | ···················· | ···················· | ···················· |
| H | 5314 | hCoV-19/Italy/LOM-UniMI-L406/2020\|EPI_ISL_542327\|2 | ···················· | ···················· | ····················· | ···················· | ···················· | ···················· |
| H | 5315 | hCoV-19/Italy/LOM-UniMI-L407/2020\|EPI_ISL_542328\|2 | ···················· | ···················· | ····················· | ···················· | ···················· | ···················· |

```
H 5316 hCoV-19/Italy/LOM-UniMI-L409/2020|EPI_ISL_542329|2 .................... .................... ..................... .................... .................... ....................
H 5317 hCoV-19/Italy/LOM-UniMI-L410/2020|EPI_ISL_542330|2 .................... .................... ..................... .................... .................... ....................
H 5318 hCoV-19/Italy/LOM-UniMI-L411/2020|EPI_ISL_542331|2 .................... .................... ..................... .................... .................... ....................
H 5319 hCoV-19/Italy/LOM-UniMI-L412/2020|EPI_ISL_542332|2 .................... .................... ..................... .................... .................... ....................
H 5320 hCoV-19/Italy/LOM-UniMI-L413/2020|EPI_ISL_542333|2 .................... .................... ..................... .................... .................... ....................
H 5321 hCoV-19/Italy/LOM-UniMI-L414/2020|EPI_ISL_542334|2 .................... .................... ..................... .................... .................... ....................
H 5322 hCoV-19/Italy/LOM-UniMI-L420/2020|EPI_ISL_542339|2 .................... .................... ..................... .................... .................... ....................
H 5324 hCoV-19/USA/TX-HMH-4415/2020|EPI_ISL_543602|2020-0 .................... .................... ..................... .................... .................... ....................
H 5327 hCoV-19/USA/TX-HMH-4419/2020|EPI_ISL_543605|2020-0 .................... .A.................. ..................... .................... .................... ....................
H 5328 hCoV-19/USA/TX-HMH-4422/2020|EPI_ISL_543606|2020-0 .................... .............A...... ..................... .................... .................... ....................
H 5329 hCoV-19/USA/TX-HMH-4423/2020|EPI_ISL_543607|2020-0 .................... .................... ..................... .................... .................... ....................
H 5330 hCoV-19/USA/TX-HMH-4424/2020|EPI_ISL_543608|2020-0 .................... .................... ..................... .................... .................... ....................
H 5331 hCoV-19/USA/TX-HMH-4425/2020|EPI_ISL_543609|2020-0 .................... .................... ..................... .................... ......T............. ....................
H 5332 hCoV-19/USA/TX-HMH-4427/2020|EPI_ISL_543611|2020-0 .................... .................... ..................... .................... .................... ....................
H 5333 hCoV-19/USA/TX-HMH-4428/2020|EPI_ISL_543612|2020-0 .................... .................... ..................... .................... .................... ....................
H 5334 hCoV-19/USA/TX-HMH-4430/2020|EPI_ISL_543615|2020-0 .................... .................... ..................... .................... .................... ....................
H 5335 hCoV-19/USA/TX-HMH-4435/2020|EPI_ISL_543619|2020-0 .................... ..........A......... ..................... .................... .................... ....................
H 5337 hCoV-19/USA/TX-HMH-4438/2020|EPI_ISL_543621|2020-0 .................... .................... ..................... .................... .................... ....................
H 5345 hCoV-19/USA/TX-HMH-4454/2020|EPI_ISL_543631|2020-0 .................... .................... ..................... .................... .................... ....................
H 5349 hCoV-19/USA/TX-HMH-4458/2020|EPI_ISL_543635|2020-0 .................... .............A...... ..................... .................... .................... ....................
H 5355 hCoV-19/USA/TX-HMH-4464/2020|EPI_ISL_543641|2020-0 .................... .................... ..................... .................... .................... ....................
H 5366 hCoV-19/USA/TX-HMH-4480/2020|EPI_ISL_543655|2020-0 .................... .................... ..................... .................... .................... ....................
H 5369 hCoV-19/USA/TX-HMH-4484/2020|EPI_ISL_543658|2020-0 .................... .................... ..................... .................... .................... ....................
H 5371 hCoV-19/USA/TX-HMH-4489/2020|EPI_ISL_543662|2020-0 .................... .................... ..................... .................... .................... ....................
H 5375 hCoV-19/USA/TX-HMH-4500/2020|EPI_ISL_543671|2020-0 .................... .................... ..................... .................... .................... ....................
H 5376 hCoV-19/USA/TX-HMH-4501/2020|EPI_ISL_543672|2020-0 .................... .................... ..................... .................... .................... ....................
H 5377 hCoV-19/England/QEUH-990682/2020|EPI_ISL_530360|20 .................... .................... ..................... .................... .................... ....................
H 5378 hCoV-19/Scotland/QEUH-9902CA/2020|EPI_ISL_530361|2 .................... .................... ..................... .................... .................... ....................
H 5379 hCoV-19/England/QEUH-99053A/2020|EPI_ISL_530362|20 .................... .................... ..................... .................... .................... ....................
H 5380 hCoV-19/Northern Ireland/QEUH-9905FE/2020|EPI_ISL_ .................... .................... ..................... .................... .................... ....................
H 5381 hCoV-19/Scotland/QEUH-9905D0/2020|EPI_ISL_530366|2 .................... .................... ..................... .................... .................... ....................
H 5382 hCoV-19/England/QEUH-9625B9/2020|EPI_ISL_531600|20 .................... .................... ..................... .................... .................... ....................
H 5383 hCoV-19/England/QEUH-9623BF/2020|EPI_ISL_531601|20 .................... .................... ..................... .................... .................... ....................
H 5384 hCoV-19/Northern Ireland/QEUH-96230A/2020|EPI_ISL_ .................... .................... ..................... .................... .................... ....................
H 5385 hCoV-19/England/QEUH-962D57/2020|EPI_ISL_531603|20 .................... .................... ..................... .................... .................... ....................
H 5386 hCoV-19/England/QEUH-963424/2020|EPI_ISL_531604|20 .................... .................... ..................... .................... .................... ....................
H 5387 hCoV-19/England/QEUH-962391/2020|EPI_ISL_531605|20 .................... .................... ..................... .................... .................... ....................
H 5388 hCoV-19/England/QEUH-962012/2020|EPI_ISL_531606|20 .................... .................... ..................... .................... .................... ....................
H 5389 hCoV-19/Scotland/QEUH-962FE8/2020|EPI_ISL_531607|2 .................... .................... ..................... .................... .................... ....................
H 5390 hCoV-19/Northern Ireland/QEUH-963257/2020|EPI_ISL_ .................... .................... ..................... .................... .................... ....................
H 5391 hCoV-19/Scotland/QEUH-963002/2020|EPI_ISL_531690|2 .................... .................... ..................... .................... .................... ....................
H 5392 hCoV-19/Scotland/QEUH-962B5D/2020|EPI_ISL_531691|2 .................... .................... ..................... .................... .................... ....................
H 5393 hCoV-19/England/QEUH-9625E6/2020|EPI_ISL_531692|20 .................... .................... ..................... .................... .................... ....................
H 5394 hCoV-19/England/QEUH-962D66/2020|EPI_ISL_531693|20 .................... .................... ..................... .................... .................... ....................
H 5395 hCoV-19/England/QEUH-96362E/2020|EPI_ISL_531694|20 .................... .................... ..................... .................... .................... ....................
H 5396 hCoV-19/England/QEUH-96265C/2020|EPI_ISL_531695|20 ........T........... .................... ..................... .................... .................... ....................
H 5397 hCoV-19/Northern Ireland/QEUH-96207C/2020|EPI_ISL_ .................... .................... ..................... .................... .................... ....................
H 5398 hCoV-19/England/QEUH-96364C/2020|EPI_ISL_531697|20 .................... .................... ..................... .................... .................... ....................
H 5399 hCoV-19/England/QEUH-7F5BD9/2020|EPI_ISL_532900|20 .................... .................... ..................... .................... .................... ....................
H 5400 hCoV-19/England/QEUH-7EC531/2020|EPI_ISL_532901|20 .................... .................... ..................... .................... .................... ....................
H 5401 hCoV-19/Northern Ireland/QEUH-7C6B77/2020|EPI_ISL_ .................... .................... ..................... .................... .................... ....................
H 5402 hCoV-19/England/QEUH-7B810D/2020|EPI_ISL_532904|20 .................... .................... ..................... .................... .................... ....................
H 5403 hCoV-19/Scotland/QEUH-7B18C3/2020|EPI_ISL_532905|2 .................... .................... ..................... .................... .................... ....................
H 5404 hCoV-19/Scotland/QEUH-7FA169/2020|EPI_ISL_532906|2 .................... .................... ..................... .................... .................... ....................
H 5405 hCoV-19/Northern Ireland/QEUH-7DA7E5/2020|EPI_ISL_ .................... .................... ..................... .................... .................... ....................
H 5406 hCoV-19/England/QEUH-7D9C41/2020|EPI_ISL_532908|20 .................... .................... ..................... .................... .................... ....................
H 5407 hCoV-19/England/QEUH-7CA0DE/2020|EPI_ISL_532911|20 .................... .................... ..................... .................... .................... ....................
H 5408 hCoV-19/England/QEUH-7C86B0/2020|EPI_ISL_532912|20 .................... .................... ..................... .................... .................... ....................
H 5409 hCoV-19/Scotland/QEUH-7AC1A2/2020|EPI_ISL_532915|2 .................... .................... ..................... .................... .................... ....................
H 5410 hCoV-19/England/QEUH-7BFF75/2020|EPI_ISL_532917|20 .................... .................... ..................... .................... .................... ....................
H 5411 hCoV-19/England/QEUH-7AB5D4/2020|EPI_ISL_532918|20 .................... .................... ..................... .................... .................... ....................
H 5412 hCoV-19/Northern Ireland/QEUH-7F0B56/2020|EPI_ISL_ .................... .................... ..................... .................... .................... ....................
H 5413 hCoV-19/England/QEUH-7B91A2/2020|EPI_ISL_532923|20 .................... .................... ..................... .................... .................... ....................
H 5414 hCoV-19/Northern Ireland/QEUH-7C82E9/2020|EPI_ISL_ .................... .................... ..................... .................... .................... ....................
H 5415 hCoV-19/England/QEUH-7EF273/2020|EPI_ISL_532927|20 .................... .................... ..................... .................... .................... ....................
H 5416 hCoV-19/England/QEUH-7BE5B0/2020|EPI_ISL_532928|20 .................... .................... ..................... .................... ..........T......... ....................
H 5417 hCoV-19/England/QEUH-7D6813/2020|EPI_ISL_532930|20 .................... .................... ..................... .................... .................... ....................
H 5418 hCoV-19/England/QEUH-7B1E49/2020|EPI_ISL_532931|20 .................... .................... ..................... .................... .................... ....................
H 5419 hCoV-19/Scotland/QEUH-7FDED8/2020|EPI_ISL_532932|2 .................... .................... ..................... .................... .................... ....................
H 5420 hCoV-19/Scotland/QEUH-7F0E4E/2020|EPI_ISL_532933|2 .................... .................... ..................... .................... .................... ....................
H 5421 hCoV-19/England/QEUH-7AFFD2/2020|EPI_ISL_532934|20 .................... .................... ..................... .................... .................... ....................
H 5422 hCoV-19/England/QEUH-7C91EB/2020|EPI_ISL_532935|20 .................... .................... ..................... .................... .................... ....................
H 5423 hCoV-19/England/QEUH-7EE8DC/2020|EPI_ISL_532938|20 .................... .................... ..................... .................... .................... ....................
H 5424 hCoV-19/England/QEUH-7C8EF4/2020|EPI_ISL_532939|20 .................... .................... ..................... .................... .................... ....................
H 5425 hCoV-19/England/OXON-B4C16/2020|EPI_ISL_535000|202 .................... .................... ..................... .................... .................... ....................
H 5426 hCoV-19/England/OXON-B4C34/2020|EPI_ISL_535001|202 .................... .................... ..................... .................... .................... ....................
H 5427 hCoV-19/England/OXON-B4C43/2020|EPI_ISL_535002|202 .................... .................... ..................... .................... .................... ....................
H 5428 hCoV-19/England/OXON-B4C8F/2020|EPI_ISL_535003|202 .................... .................... ..................... .................... .................... ....................
H 5429 hCoV-19/England/OXON-B4C9E/2020|EPI_ISL_535004|202 .................... .................... ..................... .................... .................... ....................
H 5430 hCoV-19/Scotland/CVR4046/2020|EPI_ISL_535141|2020- .................... .................... ..................... .................... .................... ....................
```

| | | | | | | | | |
|---|---|---|---|---|---|---|---|---|
| H | 5431 | hCoV-19/Scotland/CVR4050/2020\|EPI_ISL_535144\|2020- | ···················· | ···················· | ···················· | ···················· | ···················· | ···················· |
| H | 5432 | hCoV-19/Scotland/CVR4051/2020\|EPI_ISL_535145\|2020- | ···················· | ···················· | ···················· | ···················· | ···················· | ···················· |
| H | 5433 | hCoV-19/Scotland/CVR4052/2020\|EPI_ISL_535146\|2020- | ···················· | ···················· | ···················· | ···················· | ···················· | ···················· |
| H | 5434 | hCoV-19/Scotland/CVR4053/2020\|EPI_ISL_535147\|2020- | ···················· | ···················· | ···················· | ···················· | ···················· | ···················· |
| H | 5435 | hCoV-19/Scotland/CVR4055/2020\|EPI_ISL_535148\|2020- | ···················· | ···················· | ···················· | ···················· | ···················· | ···················· |
| H | 5436 | hCoV-19/Scotland/CVR4056/2020\|EPI_ISL_535149\|2020- | ···················· | ···················· | ···················· | ···················· | ···················· | ···················· |
| H | 5437 | hCoV-19/Scotland/CVR4058/2020\|EPI_ISL_535150\|2020- | ···················· | ···················· | ···················· | ···················· | ···················· | ···················· |
| H | 5438 | hCoV-19/Scotland/CVR4059/2020\|EPI_ISL_535151\|2020- | ···················· | ·A·················· | ···················· | ···················· | ···················· | ··A················· |
| H | 5439 | hCoV-19/Scotland/CVR4062/2020\|EPI_ISL_535154\|2020- | ···················· | ···················· | ···················· | ···················· | ···················· | ···················· |
| H | 5440 | hCoV-19/Scotland/CVR4064/2020\|EPI_ISL_535156\|2020- | ···················· | ···················· | ···················· | ···················· | ···················· | ···················· |
| H | 5441 | hCoV-19/Scotland/CVR4065/2020\|EPI_ISL_535157\|2020- | ···················· | ···················· | ···················· | ···················· | ···················· | ···················· |
| H | 5442 | hCoV-19/Scotland/CVR4068/2020\|EPI_ISL_535159\|2020- | ···················· | ···················· | ···················· | ···················· | ···················· | ···················· |
| H | 5443 | hCoV-19/Scotland/CVR4069/2020\|EPI_ISL_535160\|2020- | ···················· | ···················· | ···················· | ···················· | ···················· | ···················· |
| H | 5444 | hCoV-19/Scotland/CVR4071/2020\|EPI_ISL_535162\|2020- | ···················· | ·A·················· | ···················· | ···················· | ···················· | ··A················· |
| H | 5445 | hCoV-19/Scotland/CVR4073/2020\|EPI_ISL_535164\|2020- | ···················· | ···················· | ···················· | ···················· | ···················· | ···················· |
| H | 5446 | hCoV-19/Scotland/CVR4075/2020\|EPI_ISL_535166\|2020- | ···················· | ···················· | ···················· | ···················· | ···················· | ···················· |
| H | 5447 | hCoV-19/Scotland/CVR4077/2020\|EPI_ISL_535168\|2020- | ···················· | ···················· | ···················· | ···················· | ···················· | ···················· |
| H | 5448 | hCoV-19/Scotland/CVR4079/2020\|EPI_ISL_535169\|2020- | ···················· | ···················· | ···················· | ···················· | ···················· | ···················· |
| H | 5449 | hCoV-19/Scotland/CVR4081/2020\|EPI_ISL_535171\|2020- | ···················· | ·A·················· | ···················· | ···················· | ···················· | ··A················· |
| H | 5450 | hCoV-19/Scotland/CVR4082/2020\|EPI_ISL_535172\|2020- | ···················· | ···················· | ···················· | ···················· | ···················· | ···················· |
| H | 5451 | hCoV-19/Scotland/CVR4083/2020\|EPI_ISL_535173\|2020- | ···················· | ···················· | ···················· | ···················· | ···················· | ···················· |
| H | 5452 | hCoV-19/Scotland/CVR4085/2020\|EPI_ISL_535175\|2020- | ···················· | ···················· | ···················· | ···················· | ···················· | ···················· |
| H | 5453 | hCoV-19/Scotland/CVR4086/2020\|EPI_ISL_535176\|2020- | ···················· | ···················· | ···················· | ···················· | ···················· | ···················· |
| H | 5454 | hCoV-19/Scotland/CVR4087/2020\|EPI_ISL_535177\|2020- | ···················· | ·A·················· | ···················· | ···················· | ···················· | ··A················· |
| H | 5455 | hCoV-19/Scotland/CVR4088/2020\|EPI_ISL_535178\|2020- | ···················· | ···A················ | ···················· | ···················· | ···················· | ···················· |
| H | 5456 | hCoV-19/Scotland/CVR4090/2020\|EPI_ISL_535180\|2020- | ···················· | ···················· | ···················· | ···················· | ···················· | ···················· |
| H | 5457 | hCoV-19/Scotland/CVR4092/2020\|EPI_ISL_535182\|2020- | ···················· | ···················· | ···················· | ···················· | ···················· | ···················· |
| H | 5458 | hCoV-19/Wales/PHWC-169FDA/2020\|EPI_ISL_535184\|2020 | ···················· | ···················· | ···················· | ···················· | ···················· | ···················· |
| H | 5459 | hCoV-19/Wales/PHWC-169FE9/2020\|EPI_ISL_535185\|2020 | ···················· | ···················· | ···················· | ···················· | ···················· | ···················· |
| H | 5460 | hCoV-19/Wales/PHWC-169FF8/2020\|EPI_ISL_535186\|2020 | ···················· | ···················· | ···················· | ···················· | ···················· | ···················· |
| H | 5461 | hCoV-19/Wales/PHWC-16A012/2020\|EPI_ISL_535188\|2020 | ···················· | ···················· | ···················· | ···················· | ···················· | ···················· |
| H | 5462 | hCoV-19/Wales/PHWC-16A021/2020\|EPI_ISL_535189\|2020 | ···················· | ···················· | ···················· | ···················· | ···················· | ···················· |
| H | 5463 | hCoV-19/Wales/PHWC-16A030/2020\|EPI_ISL_535190\|2020 | ···················· | ···················· | ···················· | ···················· | ···················· | ···················· |
| H | 5464 | hCoV-19/Wales/PHWC-16A04F/2020\|EPI_ISL_535191\|2020 | ···················· | ···················· | ···················· | ···················· | ···················· | ···················· |
| H | 5465 | hCoV-19/Wales/PHWC-16A05E/2020\|EPI_ISL_535192\|2020 | ···················· | ···················· | ···················· | ···················· | ···················· | ···················· |
| H | 5466 | hCoV-19/Wales/PHWC-16A06D/2020\|EPI_ISL_535193\|2020 | ···················· | ···················· | ···················· | ···················· | ···················· | ···················· |
| H | 5467 | hCoV-19/Wales/PHWC-16A07C/2020\|EPI_ISL_535194\|2020 | ···················· | ···················· | ···················· | ···················· | ···················· | ···················· |
| H | 5468 | hCoV-19/Wales/PHWC-16A08B/2020\|EPI_ISL_535195\|2020 | ···················· | ···················· | ···················· | ···················· | ···················· | ···················· |
| H | 5469 | hCoV-19/Wales/PHWC-16A09A/2020\|EPI_ISL_535196\|2020 | ···················· | ···················· | ···················· | ···················· | ···················· | ···················· |
| H | 5470 | hCoV-19/Wales/PHWC-16A0A9/2020\|EPI_ISL_535197\|2020 | ···················· | ···················· | ···················· | ···················· | ···················· | ···················· |
| H | 5471 | hCoV-19/Wales/PHWC-16A0B8/2020\|EPI_ISL_535198\|2020 | ···················· | ···················· | ···················· | ···················· | ···················· | ···················· |
| H | 5472 | hCoV-19/Wales/PHWC-16A0D6/2020\|EPI_ISL_535199\|2020 | ···················· | ···················· | ···················· | ···················· | ···················· | ···················· |
| H | 5473 | hCoV-19/Canada/QC-LSPQ-L00241965/2020\|EPI_ISL_5363 | ···················· | ···················· | ···················· | ···················· | ···················· | ···················· |
| H | 5474 | hCoV-19/Canada/QC-LSPQ-L00241993/2020\|EPI_ISL_5363 | ···················· | ···················· | ···················· | ···················· | ···················· | ···················· |
| H | 5475 | hCoV-19/Canada/QC-LSPQ-L00242011/2020\|EPI_ISL_5363 | ···················· | ···················· | ···················· | ···················· | ···················· | ···················· |
| H | 5476 | hCoV-19/Canada/QC-LSPQ-L00242018/2020\|EPI_ISL_5363 | ···················· | ···················· | ···················· | ···················· | ···················· | ···················· |
| H | 5477 | hCoV-19/Canada/QC-LSPQ-L00242019/2020\|EPI_ISL_5363 | ···················· | ···················· | ···················· | ···················· | ···················· | ···················· |
| H | 5478 | hCoV-19/Canada/QC-LSPQ-L00242033/2020\|EPI_ISL_5363 | ···················· | ···················· | ···················· | ···················· | ···················· | ···················· |
| H | 5479 | hCoV-19/Canada/QC-LSPQ-L00242035/2020\|EPI_ISL_5363 | ···················· | ···················· | ···················· | ···················· | ···················· | ···················· |
| H | 5480 | hCoV-19/Canada/QC-LSPQ-L00242051/2020\|EPI_ISL_5363 | ···················· | ···················· | ···················· | ···················· | ···················· | ···················· |
| H | 5481 | hCoV-19/Canada/QC-LSPQ-L00242105/2020\|EPI_ISL_5363 | ···················· | ···················· | ···················· | ···················· | ···················· | ···················· |
| H | 5482 | hCoV-19/Canada/QC-LSPQ-L00242228/2020\|EPI_ISL_5363 | ···················· | ···················· | ···················· | ···················· | ···················· | ···················· |
| H | 5483 | hCoV-19/Canada/QC-LSPQ-L00242235/2020\|EPI_ISL_5363 | ···················· | ···················· | ···················· | ···················· | ···················· | ···················· |
| H | 5484 | hCoV-19/Canada/QC-LSPQ-L00242238/2020\|EPI_ISL_5363 | ···················· | ···················· | ···················· | ···················· | ···················· | ···················· |
| H | 5485 | hCoV-19/Canada/QC-LSPQ-L00242384/2020\|EPI_ISL_5363 | ···················· | ···················· | ···················· | ···················· | ···················· | ···················· |
| H | 5486 | hCoV-19/Canada/QC-LSPQ-L00242487/2020\|EPI_ISL_5363 | ···················· | ···················· | ···················· | ···················· | ···················· | ···················· |
| H | 5487 | hCoV-19/Canada/QC-LSPQ-L00242505/2020\|EPI_ISL_5363 | ···················· | ···················· | ···················· | ···················· | ···················· | ···················· |
| H | 5488 | hCoV-19/Canada/QC-LSPQ-L00242547/2020\|EPI_ISL_5363 | ···················· | ···················· | ···················· | ···················· | ···················· | ···················· |
| H | 5489 | hCoV-19/Canada/QC-LSPQ-L00242576/2020\|EPI_ISL_5363 | ···················· | ···················· | ···················· | ···················· | ···················· | ···················· |
| H | 5490 | hCoV-19/Canada/QC-LSPQ-L00242662/2020\|EPI_ISL_5363 | ···················· | ···················· | ···················· | ···················· | ···················· | ···················· |
| H | 5491 | hCoV-19/Canada/QC-LSPQ-L00242684/2020\|EPI_ISL_5363 | ···················· | ···················· | ···················· | ···················· | ···················· | ···················· |
| H | 5492 | hCoV-19/Canada/QC-LSPQ-L00242714/2020\|EPI_ISL_5363 | ···················· | ···················· | ···················· | ···················· | ···················· | ···················· |
| H | 5493 | hCoV-19/Peru/LIM-INS-077/2020\|EPI_ISL_536477\|2020- | ···················· | ···················· | ···················· | ···················· | ···················· | ···················· |
| H | 5494 | hCoV-19/Peru/LIM-INS-078/2020\|EPI_ISL_536478\|2020- | ···················· | ···················· | ···················· | ···················· | ···················· | ···················· |
| H | 5495 | hCoV-19/Peru/CAL-INS-079/2020\|EPI_ISL_536479\|2020- | ···················· | ···················· | ···················· | ···················· | ···················· | ···················· |
| H | 5496 | hCoV-19/Peru/CAL-INS-081/2020\|EPI_ISL_536480\|2020- | ···················· | ···················· | ···················· | ···················· | ···················· | ···················· |
| H | 5497 | hCoV-19/Peru/CAL-INS-082/2020\|EPI_ISL_536481\|2020- | ···················· | ···················· | ···················· | ···················· | ···················· | ···················· |
| H | 5498 | hCoV-19/Peru/ANC-INS-083/2020\|EPI_ISL_536482\|2020- | ···················· | ···················· | ···················· | ···················· | ···················· | ···················· |
| H | 5499 | hCoV-19/Peru/ANC-INS-084/2020\|EPI_ISL_536483\|2020- | ···················· | ···················· | ···················· | ···················· | ···················· | ···················· |
| H | 5500 | hCoV-19/Peru/ANC-INS-085/2020\|EPI_ISL_536484\|2020- | ···················· | ···················· | ···················· | ···················· | ···················· | ···················· |
| H | 5501 | hCoV-19/Peru/LIM-INS-086/2020\|EPI_ISL_536485\|2020- | ···················· | ···················· | ···················· | ···················· | ···················· | ···················· |
| H | 5502 | hCoV-19/Peru/LIM-INS-087/2020\|EPI_ISL_536486\|2020- | ···················· | ···················· | ···················· | ···················· | ···················· | ···················· |
| H | 5503 | hCoV-19/Peru/LIM-INS-088/2020\|EPI_ISL_536487\|2020- | ···················· | ···················· | ···················· | ···················· | ···················· | ···················· |
| H | 5504 | hCoV-19/Peru/LIM-INS-089/2020\|EPI_ISL_536488\|2020- | ···················· | ···················· | ···················· | ···················· | ···················· | ···················· |
| H | 5505 | hCoV-19/Peru/LIM-INS-090/2020\|EPI_ISL_536489\|2020- | ···················· | ···················· | ···················· | ···················· | ···················· | ···················· |
| H | 5506 | hCoV-19/Peru/LIM-INS-091/2020\|EPI_ISL_536490\|2020- | ···················· | ···················· | ···················· | ···················· | ···················· | ···················· |
| H | 5507 | hCoV-19/Peru/ANC-INS-092/2020\|EPI_ISL_536491\|2020- | ···················· | ···················· | ···················· | ···················· | ···················· | ···················· |
| H | 5508 | hCoV-19/Peru/LIM-INS-093/2020\|EPI_ISL_536492\|2020- | ···················· | ···················· | ···················· | ···················· | ···················· | ···················· |
| H | 5509 | hCoV-19/Peru/LIM-INS-094/2020\|EPI_ISL_536493\|2020- | ···················· | ···················· | ···················· | ···················· | ···················· | ···················· |
| H | 5510 | hCoV-19/Peru/LIM-INS-095/2020\|EPI_ISL_536494\|2020- | ···················· | ···················· | ···················· | ···················· | ···················· | ···················· |

| | | | | | | | | |
|---|---|---|---|---|---|---|---|---|
| H | 5511 | hCoV-19/Peru/LIM-INS-096/2020\|EPI_ISL_536495\|2020- | ···················· | ···················· | ····················· | ···················· | ···················· | ···················· |
| H | 5512 | hCoV-19/Peru/LIM-INS-097/2020\|EPI_ISL_536496\|2020- | ···················· | ···················· | ····················· | ···················· | ···················· | ···················· |
| H | 5513 | hCoV-19/Peru/LIM-INS-099/2020\|EPI_ISL_536498\|2020- | ···················· | ···················· | ····················· | ···················· | ···················· | ···················· |
| H | 5514 | hCoV-19/Peru/LIM-INS-100/2020\|EPI_ISL_536499\|2020- | ···················· | ···················· | ····················· | ···················· | ···················· | ···················· |
| H | 5515 | hCoV-19/USA/CA-UCLA-81/2020\|EPI_ISL_537602\|2020-03 | ···················· | ····A··············· | ····················· | ···················· | ···················· | ···················· |
| H | 5516 | hCoV-19/USA/CA-UCLA-84/2020\|EPI_ISL_537603\|2020-03 | ···················· | ···················· | ····················· | ···················· | ···················· | ···················· |
| H | 5517 | hCoV-19/USA/CA-UCLA-87/2020\|EPI_ISL_537604\|2020-03 | ···················· | ···················· | ····················· | ···················· | ···················· | ···················· |
| H | 5518 | hCoV-19/USA/CA-UCLA-9/2020\|EPI_ISL_537605\|2020-04- | ···················· | ···················· | ····················· | ···················· | ···················· | ···················· |
| H | 5519 | hCoV-19/USA/CA-UCLA-97/2020\|EPI_ISL_537606\|2020-03 | ···················· | ···················· | ····················· | ···················· | ···················· | ···················· |
| H | 5520 | hCoV-19/USA/CA-UCLA-98/2020\|EPI_ISL_537607\|2020-03 | ···················· | ···················· | ····················· | ···················· | ···················· | ···················· |
| H | 5535 | hCoV-19/England/LEED-2AA455/2020\|EPI_ISL_538925\|20 | ···················· | ···················· | ····················· | ···················· | ···················· | ···················· |
| H | 5536 | hCoV-19/England/LEED-2AA25B/2020\|EPI_ISL_538926\|20 | ···················· | ···················· | ····················· | ···················· | ···················· | ···················· |
| H | 5537 | hCoV-19/England/LEED-2A9CF4/2020\|EPI_ISL_538928\|20 | ···················· | ···················· | ····················· | ···················· | ···················· | ···················· |
| H | 5538 | hCoV-19/England/LEED-2A9C7C/2020\|EPI_ISL_538929\|20 | ···················· | ·A·················· | ····················· | ···················· | ······K············· | ···················· |
| H | 5539 | hCoV-19/England/LEED-2A94A1/2020\|EPI_ISL_538930\|20 | ···················· | ···················· | ····················· | ···················· | ···················· | ···················· |
| H | 5540 | hCoV-19/England/LEED-2AA1E5/2020\|EPI_ISL_538931\|20 | ···················· | ···················· | ····················· | ···················· | ···················· | ···················· |
| H | 5541 | hCoV-19/England/LEED-2AA1F4/2020\|EPI_ISL_538932\|20 | ···················· | ···················· | ····················· | ···················· | ···················· | ···················· |
| H | 5542 | hCoV-19/England/LEED-2AA130/2020\|EPI_ISL_538933\|20 | ···················· | ···················· | ····················· | ···················· | ···················· | ···················· |
| H | 5543 | hCoV-19/England/LEED-2A9E76/2020\|EPI_ISL_538934\|20 | ···················· | ···················· | ····················· | ···················· | ···················· | ···················· |
| H | 5544 | hCoV-19/England/LEED-2A9E58/2020\|EPI_ISL_538935\|20 | ···················· | ···················· | ····················· | ···················· | ···················· | ···················· |
| H | 5545 | hCoV-19/England/LEED-2A9C03/2020\|EPI_ISL_538936\|20 | ···················· | ···················· | ····················· | ···················· | ···················· | ···················· |
| H | 5546 | hCoV-19/England/LEED-2A9B60/2020\|EPI_ISL_538937\|20 | ···················· | ···················· | ····················· | ···················· | ···················· | ···················· |
| H | 5547 | hCoV-19/England/LEED-2A9E3A/2020\|EPI_ISL_538938\|20 | ···················· | ···················· | ····················· | ···················· | ···················· | ···················· |
| H | 5548 | hCoV-19/England/LEED-2A9DC4/2020\|EPI_ISL_538939\|20 | ···················· | ·A·················· | ····················· | ···················· | ···················· | ···················· |
| H | 5549 | hCoV-19/England/LEED-2A9CB8/2020\|EPI_ISL_538940\|20 | ···················· | ···················· | ····················· | ···················· | ···················· | ···················· |
| H | 5550 | hCoV-19/England/LEED-2A9D88/2020\|EPI_ISL_538941\|20 | ···················· | ···················· | ····················· | ···················· | ···················· | ···················· |
| H | 5551 | hCoV-19/England/LEED-2A9CA9/2020\|EPI_ISL_538942\|20 | ···················· | ·A·················· | ····················· | ···················· | ···················· | ···················· |
| H | 5552 | hCoV-19/England/LEED-2A9FEC/2020\|EPI_ISL_538943\|20 | ···················· | ···················· | ····················· | ···················· | ···················· | ···················· |
| H | 5553 | hCoV-19/England/LEED-2AA798/2020\|EPI_ISL_538944\|20 | ···················· | ···················· | ····················· | ···················· | ···················· | ···················· |
| H | 5554 | hCoV-19/England/LEED-2A9C30/2020\|EPI_ISL_538946\|20 | ···················· | ···················· | ····················· | ···················· | ···················· | ···················· |
| H | 5555 | hCoV-19/England/LEED-2A9CC7/2020\|EPI_ISL_538947\|20 | ···················· | ···················· | ····················· | ···················· | ···················· | ···················· |
| H | 5556 | hCoV-19/England/LEED-2A9D3D/2020\|EPI_ISL_538948\|20 | ···················· | ···················· | ····················· | ···················· | ···················· | ···················· |
| H | 5557 | hCoV-19/England/LEED-2AA042/2020\|EPI_ISL_538949\|20 | ···················· | ······R············· | ····················· | ···················· | ···················· | ···················· |
| H | 5558 | hCoV-19/England/LEED-2A9D97/2020\|EPI_ISL_538950\|20 | ···················· | ···················· | ····················· | ···················· | ···················· | ···················· |
| H | 5559 | hCoV-19/England/LEED-2A9CD6/2020\|EPI_ISL_538951\|20 | ···················· | ···················· | ····················· | ···················· | ···················· | ···················· |
| H | 5560 | hCoV-19/England/LEED-2AA76B/2020\|EPI_ISL_538952\|20 | ···················· | ···················· | ····················· | ···················· | ···················· | ···················· |
| H | 5561 | hCoV-19/England/LEED-2AA6AA/2020\|EPI_ISL_538953\|20 | ···················· | ···················· | ····················· | ···················· | ···················· | ···················· |
| H | 5562 | hCoV-19/England/LEED-2A9D79/2020\|EPI_ISL_538954\|20 | ···················· | ···················· | ····················· | ···················· | ···················· | ···················· |
| H | 5563 | hCoV-19/England/LEED-2A9CE5/2020\|EPI_ISL_538955\|20 | ···················· | ···················· | ····················· | ···················· | ···················· | ···················· |
| H | 5564 | hCoV-19/England/LEED-2A9DF1/2020\|EPI_ISL_538956\|20 | ···················· | ···················· | ····················· | ···················· | ···················· | ···················· |
| H | 5565 | hCoV-19/England/LEED-2A9EEF/2020\|EPI_ISL_538957\|20 | ···················· | ···················· | ····················· | ···················· | ···················· | ···················· |
| H | 5566 | hCoV-19/England/LEED-2AA75C/2020\|EPI_ISL_538958\|20 | ···················· | ···················· | ····················· | ···················· | ···················· | ···················· |
| H | 5567 | hCoV-19/England/LEED-2A9F55/2020\|EPI_ISL_538959\|20 | ···················· | ···················· | ····················· | ···················· | ···················· | ···················· |
| H | 5568 | hCoV-19/England/LEED-2A9BF7/2020\|EPI_ISL_538960\|20 | ···················· | ·A·················· | ····················· | ···················· | ···················· | ···················· |
| H | 5569 | hCoV-19/England/LEED-2A9E1C/2020\|EPI_ISL_538961\|20 | ···················· | ···················· | ····················· | ···················· | ···················· | ···················· |
| H | 5570 | hCoV-19/England/LEED-2A9D6A/2020\|EPI_ISL_538962\|20 | ···················· | ·A·················· | ····················· | ···················· | ···················· | ···················· |
| H | 5571 | hCoV-19/England/LEED-2A9DA6/2020\|EPI_ISL_538963\|20 | ···················· | ···················· | ····················· | ···················· | ···················· | ···················· |
| H | 5572 | hCoV-19/England/LEED-2A9E0D/2020\|EPI_ISL_538965\|20 | ···················· | ···················· | ····················· | ···················· | ···················· | ···················· |
| H | 5573 | hCoV-19/England/LEED-2A9D4C/2020\|EPI_ISL_538966\|20 | ···················· | ···················· | ····················· | ···················· | ···················· | ···················· |
| H | 5574 | hCoV-19/England/LEED-2AA033/2020\|EPI_ISL_538967\|20 | ···················· | ···················· | ····················· | ···················· | ···················· | ···················· |
| H | 5575 | hCoV-19/England/LEED-2A9D1F/2020\|EPI_ISL_538968\|20 | ···················· | ···················· | ····················· | ···················· | ···················· | ···················· |
| H | 5576 | hCoV-19/England/LEED-2AA789/2020\|EPI_ISL_538969\|20 | ···················· | ·A·················· | ····················· | ···················· | ···················· | ···················· |
| H | 5577 | hCoV-19/England/LEED-2AA7A7/2020\|EPI_ISL_538970\|20 | ···················· | ···················· | ····················· | ···················· | ···················· | ···················· |
| H | 5578 | hCoV-19/England/LEED-2A9E85/2020\|EPI_ISL_538971\|20 | ···················· | ···················· | ····················· | ···················· | ···················· | ···················· |
| H | 5579 | hCoV-19/England/LEED-2A9D2E/2020\|EPI_ISL_538972\|20 | ···················· | ···················· | ····················· | ···················· | ···················· | ···················· |
| H | 5580 | hCoV-19/England/LEED-2A9ED0/2020\|EPI_ISL_538973\|20 | ···················· | ·A·················· | ····················· | ···················· | ···················· | ···················· |
| H | 5581 | hCoV-19/England/LEED-2A9E49/2020\|EPI_ISL_538974\|20 | ···················· | ···················· | ····················· | ···················· | ···················· | ···················· |
| H | 5582 | hCoV-19/England/LEED-2A9E67/2020\|EPI_ISL_538975\|20 | ···················· | ···················· | ····················· | ···················· | ···················· | ···················· |
| H | 5583 | hCoV-19/England/LEED-2A9C9A/2020\|EPI_ISL_538976\|20 | ···················· | ···················· | ····················· | ···················· | ···················· | ···················· |
| H | 5584 | hCoV-19/England/LEED-2A9F64/2020\|EPI_ISL_538977\|20 | ···················· | ···················· | ····················· | ···················· | ···················· | ···················· |
| H | 5585 | hCoV-19/England/LEED-2AA72F/2020\|EPI_ISL_538978\|20 | ···················· | ·A·················· | ····················· | ···················· | ···················· | ···················· |
| H | 5586 | hCoV-19/England/LEED-2AA6F5/2020\|EPI_ISL_538979\|20 | ···················· | ···················· | ····················· | ···················· | ···················· | ···················· |
| H | 5587 | hCoV-19/England/LEED-2AA701/2020\|EPI_ISL_538980\|20 | ···················· | ···················· | ····················· | ···················· | ···················· | ···················· |
| H | 5588 | hCoV-19/England/LEED-2AA6B9/2020\|EPI_ISL_538981\|20 | ···················· | ···················· | ····················· | ···················· | ···················· | ···················· |
| H | 5589 | hCoV-19/England/LEED-2AA6D7/2020\|EPI_ISL_538982\|20 | ···················· | ···················· | ····················· | ···················· | ···················· | ···················· |
| H | 5590 | hCoV-19/England/LEED-2AA6E6/2020\|EPI_ISL_538983\|20 | ···················· | ···················· | ····················· | ···················· | ···················· | ···················· |
| H | 5591 | hCoV-19/England/LEED-2AA6C8/2020\|EPI_ISL_538984\|20 | ···················· | ···················· | ····················· | ···················· | ···················· | ···················· |
| H | 5592 | hCoV-19/England/LEED-2AA710/2020\|EPI_ISL_538985\|20 | ···················· | ···················· | ····················· | ···················· | ···················· | ···················· |
| H | 5593 | hCoV-19/England/LEED-2A832D/2020\|EPI_ISL_538986\|20 | ···················· | ···················· | ····················· | ···················· | ···················· | ···················· |
| H | 5594 | hCoV-19/England/LEED-2A830F/2020\|EPI_ISL_538988\|20 | ···················· | ···················· | ····················· | ···················· | ···················· | ···················· |
| H | 5595 | hCoV-19/England/LEED-2A82D5/2020\|EPI_ISL_538989\|20 | ···················· | ····A··············· | ····················· | ···················· | ···················· | ···················· |
| H | 5596 | hCoV-19/England/LEED-2A828A/2020\|EPI_ISL_538990\|20 | ···················· | ···················· | ····················· | ···················· | ···················· | ···················· |
| H | 5597 | hCoV-19/England/LEED-2A83F0/2020\|EPI_ISL_538991\|20 | ···················· | ···················· | ····················· | ···················· | ···················· | ···················· |
| H | 5598 | hCoV-19/England/LEED-2A8211/2020\|EPI_ISL_538992\|20 | ···················· | ···················· | ····················· | ···················· | ···················· | ···················· |
| H | 5599 | hCoV-19/England/LEED-2A83C3/2020\|EPI_ISL_538993\|20 | ···················· | ····M··············· | ····················· | ···················· | ···················· | ···················· |
| H | 5600 | hCoV-19/England/LEED-2A83D2/2020\|EPI_ISL_538998\|20 | ···················· | ···················· | ····················· | ···················· | ···················· | ···················· |
| H | 5601 | hCoV-19/England/LEED-2A8202/2020\|EPI_ISL_538999\|20 | ···················· | ···················· | ····················· | ···················· | ···················· | ···················· |
| H | 5602 | hCoV-19/South Korea/IHC18561/2020\|EPI_ISL_541004\|2 | ···················· | ···················· | ····················· | ···················· | ···················· | ···················· |
| H | 5603 | hCoV-19/South Korea/IHC18670/2020\|EPI_ISL_541005\|2 | ···················· | ···················· | ····················· | ···················· | ···················· | ···················· |
| H | 5604 | hCoV-19/South Korea/IHC18755/2020\|EPI_ISL_541006\|2 | ···················· | ···················· | ····················· | ···················· | ···················· | ···················· |

| | | | | | | | | |
|---|---|---|---|---|---|---|---|---|
| H | 5605 | hCoV-19/South Korea/IHC20065/2020\|EPI_ISL_541007\|2 | ···················· | ···················· | ···················· | ···················· | ···················· | ···················· |
| H | 5606 | hCoV-19/Italy/LOM-UniMI-L201/2020\|EPI_ISL_542190\|2 | ···················· | ···················· | ···················· | ···················· | ···················· | ···················· |
| H | 5607 | hCoV-19/Italy/LOM-UniMI-L203/2020\|EPI_ISL_542191\|2 | ···················· | ···················· | ···················· | ···················· | ···················· | ···················· |
| H | 5608 | hCoV-19/Italy/LOM-UniMI-L204/2020\|EPI_ISL_542192\|2 | ···················· | ···················· | ···················· | ···················· | ···················· | ···················· |
| H | 5609 | hCoV-19/Italy/LOM-UniMI-L205/2020\|EPI_ISL_542193\|2 | ···················· | ···················· | ···················· | ···················· | ···················· | ···················· |
| H | 5610 | hCoV-19/Italy/LOM-UniMI-L206/2020\|EPI_ISL_542194\|2 | ···················· | ···················· | ···················· | ···················· | ···················· | ···················· |
| H | 5611 | hCoV-19/Italy/LOM-UniMI-L207/2020\|EPI_ISL_542195\|2 | ···················· | ···················· | ···················· | ···················· | ···················· | ···················· |
| H | 5612 | hCoV-19/Italy/LOM-UniMI-L208/2020\|EPI_ISL_542196\|2 | ···················· | ···················· | ···················· | ···················· | ···················· | ···················· |
| H | 5613 | hCoV-19/Italy/LOM-UniMI-L210/2020\|EPI_ISL_542197\|2 | ···················· | ···················· | ···················· | ···················· | ···················· | ···················· |
| H | 5614 | hCoV-19/Italy/LOM-UniMI-L211/2020\|EPI_ISL_542198\|2 | ···················· | ···················· | ···················· | ···················· | ···················· | ···················· |
| H | 5615 | hCoV-19/Italy/LOM-UniMI-L212/2020\|EPI_ISL_542199\|2 | ···················· | ···················· | ···················· | ···················· | ···················· | ···················· |
| H | 5616 | hCoV-19/Italy/LOM-UniMI-L415/2020\|EPI_ISL_542335\|2 | ···················· | ···················· | ···················· | ···················· | ···················· | ···················· |
| H | 5617 | hCoV-19/Italy/LOM-UniMI-L416/2020\|EPI_ISL_542336\|2 | ···················· | ···················· | ···················· | ···················· | ···················· | ···················· |
| H | 5618 | hCoV-19/Italy/LOM-UniMI-L418/2020\|EPI_ISL_542337\|2 | ···················· | ···················· | ···················· | ···················· | ···················· | ···················· |
| H | 5619 | hCoV-19/Italy/LOM-UniMI-L419/2020\|EPI_ISL_542338\|2 | ···················· | ···················· | ···················· | ···················· | ···················· | ···················· |
| H | 5620 | hCoV-19/Italy/LOM-UniMI-L421/2020\|EPI_ISL_542340\|2 | ···················· | ···················· | ···················· | ···················· | ···················· | ···················· |
| H | 5621 | hCoV-19/Italy/LOM-UniMI-L422/2020\|EPI_ISL_542341\|2 | ···················· | ···················· | ···················· | ···················· | ···················· | ···················· |
| H | 5622 | hCoV-19/Italy/LOM-UniMI-L423/2020\|EPI_ISL_542342\|2 | ···················· | ···················· | ···················· | ···················· | ···················· | ···················· |
| H | 5623 | hCoV-19/Italy/LOM-UniMI-L424/2020\|EPI_ISL_542343\|2 | ···················· | ···················· | ···················· | ···················· | ···················· | ···················· |
| H | 5624 | hCoV-19/Italy/LOM-UniMI-L425/2020\|EPI_ISL_542344\|2 | ···················· | ···················· | ···················· | ···················· | ···················· | ···················· |
| H | 5625 | hCoV-19/Italy/LOM-UniMI-L426/2020\|EPI_ISL_542345\|2 | ···················· | ···················· | ···················· | ···················· | ···················· | ···················· |
| H | 5628 | hCoV-19/USA/TX-HMH-4497/2020\|EPI_ISL_543668\|2020-0 | ···················· | ···················· | ···················· | ···················· | ···················· | ···················· |
| H | 5629 | hCoV-19/USA/TX-HMH-4502/2020\|EPI_ISL_543673\|2020-0 | ···················· | ···················· | ···················· | ···················· | ···················· | ···················· |
| H | 5630 | hCoV-19/USA/TX-HMH-4504/2020\|EPI_ISL_543674\|2020-0 | ···················· | ···················· | ···················· | ···················· | ···················· | ···················· |
| H | 5633 | hCoV-19/Northern Ireland/QEUH-96AB64/2020\|EPI_ISL_ | ···················· | ···················· | ···················· | ···················· | ···················· | ···················· |
| H | 5634 | hCoV-19/England/QEUH-96B35C/2020\|EPI_ISL_531457\|20 | ···················· | ···················· | ···················· | ···················· | ···················· | ···················· |
| H | 5635 | hCoV-19/England/QEUH-96B1CB/2020\|EPI_ISL_531458\|20 | ···················· | ···················· | ···················· | ···················· | ···················· | ···················· |
| H | 5636 | hCoV-19/England/QEUH-96BCE8/2020\|EPI_ISL_531459\|20 | ···················· | ···················· | ···················· | ···················· | ···················· | ···················· |
| H | 5637 | hCoV-19/Northern Ireland/QEUH-96B92D/2020\|EPI_ISL_ | ···················· | ···················· | ···················· | ···················· | ···················· | ···················· |
| H | 5638 | hCoV-19/England/QEUH-96B468/2020\|EPI_ISL_531462\|20 | ···················· | ···················· | ···················· | ···················· | ···················· | ···················· |
| H | 5639 | hCoV-19/England/QEUH-96A690/2020\|EPI_ISL_531463\|20 | ···················· | ···················· | ···················· | ···················· | ···················· | ···················· |
| H | 5640 | hCoV-19/Scotland/QEUH-9630D5/2020\|EPI_ISL_531464\|2 | ···················· | ···················· | ···················· | ···················· | ···················· | ···················· |
| H | 5641 | hCoV-19/England/QEUH-96A487/2020\|EPI_ISL_531465\|20 | ···················· | ···················· | ···················· | ···················· | ···················· | ···················· |
| H | 5642 | hCoV-19/England/QEUH-96A821/2020\|EPI_ISL_531466\|20 | ···················· | ···················· | ···················· | ···················· | ···················· | ···················· |
| H | 5643 | hCoV-19/England/QEUH-96A7D9/2020\|EPI_ISL_531467\|20 | ···················· | ···················· | ···················· | ···················· | ···················· | ···················· |
| H | 5644 | hCoV-19/England/QEUH-96A40F/2020\|EPI_ISL_531468\|20 | ···················· | ···················· | ···················· | ···················· | ···················· | ···················· |
| H | 5645 | hCoV-19/England/QEUH-96A4D2/2020\|EPI_ISL_531469\|20 | ···················· | ···················· | ···················· | ···················· | ···················· | ···················· |
| H | 5646 | hCoV-19/Scotland/QEUH-96A7AC/2020\|EPI_ISL_531470\|2 | ···················· | ···················· | ···················· | ···················· | ···················· | ···················· |
| H | 5647 | hCoV-19/England/QEUH-963C77/2020\|EPI_ISL_531471\|20 | ···················· | ···················· | ···················· | ···················· | ···················· | ···················· |
| H | 5648 | hCoV-19/England/QEUH-963D0B/2020\|EPI_ISL_531472\|20 | ···················· | ···················· | ···················· | ···················· | ···················· | ···················· |
| H | 5649 | hCoV-19/Scotland/QEUH-9627E0/2020\|EPI_ISL_531473\|2 | ···················· | ···················· | ···················· | ···················· | ···················· | ···················· |
| H | 5650 | hCoV-19/England/QEUH-96A681/2020\|EPI_ISL_531475\|20 | ···················· | ···················· | ···················· | ···················· | ···················· | ···················· |
| H | 5651 | hCoV-19/England/QEUH-963A13/2020\|EPI_ISL_531479\|20 | ···················· | ···················· | ···················· | ···················· | ···················· | ···················· |
| H | 5652 | hCoV-19/Scotland/QEUH-96A751/2020\|EPI_ISL_531480\|2 | ···················· | ···················· | ···················· | ···················· | ···················· | ···················· |
| H | 5653 | hCoV-19/England/QEUH-962874/2020\|EPI_ISL_531482\|20 | ···················· | ···················· | ···················· | ···················· | ···················· | ···················· |
| H | 5654 | hCoV-19/Scotland/QEUH-96313C/2020\|EPI_ISL_531483\|2 | ···················· | ···················· | ···················· | ···················· | ···················· | ···················· |
| H | 5655 | hCoV-19/Scotland/QEUH-963DFC/2020\|EPI_ISL_531484\|2 | ···················· | ···················· | ···················· | ···················· | ···················· | ···················· |
| H | 5656 | hCoV-19/Scotland/QEUH-96A6DC/2020\|EPI_ISL_531485\|2 | ···················· | ···················· | ···················· | ···················· | ···················· | ···················· |
| H | 5657 | hCoV-19/England/QEUH-963E71/2020\|EPI_ISL_531486\|20 | ···················· | ···················· | ···················· | ···················· | ···················· | ···················· |
| H | 5658 | hCoV-19/England/QEUH-963D29/2020\|EPI_ISL_531487\|20 | ···················· | ···················· | ···················· | ···················· | ···················· | ···················· |
| H | 5659 | hCoV-19/England/QEUH-963B7A/2020\|EPI_ISL_531489\|20 | ···················· | ···················· | ···················· | ···················· | ···················· | ···················· |
| H | 5660 | hCoV-19/England/QEUH-962786/2020\|EPI_ISL_531490\|20 | ···················· | ···················· | ···················· | ···················· | ···················· | ···················· |
| H | 5661 | hCoV-19/England/QEUH-962829/2020\|EPI_ISL_531491\|20 | ···················· | ···················· | ···················· | ···················· | ···················· | ···················· |
| H | 5662 | hCoV-19/Scotland/QEUH-9630F3/2020\|EPI_ISL_531492\|2 | ···················· | ···················· | ···················· | ···················· | ···················· | ···················· |
| H | 5663 | hCoV-19/England/QEUH-963BA7/2020\|EPI_ISL_531494\|20 | ···················· | ···················· | ···················· | ···················· | ···················· | ···················· |
| H | 5664 | hCoV-19/Scotland/QEUH-96A6CD/2020\|EPI_ISL_531495\|2 | ···················· | ···················· | ···················· | ···················· | ···················· | ···················· |
| H | 5665 | hCoV-19/England/QEUH-963AAA/2020\|EPI_ISL_531496\|20 | ···················· | ···················· | ···················· | ···················· | ···················· | ···················· |
| H | 5666 | hCoV-19/England/QEUH-96A645/2020\|EPI_ISL_531497\|20 | ···················· | ···················· | ···················· | ···················· | ···················· | ···················· |
| H | 5667 | hCoV-19/England/QEUH-96A715/2020\|EPI_ISL_531498\|20 | ···················· | ···················· | ···················· | ···················· | ···················· | ···················· |
| H | 5668 | hCoV-19/England/QEUH-963A5F/2020\|EPI_ISL_531499\|20 | ···················· | ···················· | ···················· | ···················· | ···················· | ···················· |
| H | 5669 | hCoV-19/Scotland/QEUH-9627D1/2020\|EPI_ISL_531500\|2 | ···················· | ·A·················· | ···················· | ···················· | ···················· | ···················· |
| H | 5670 | hCoV-19/England/QEUH-96A618/2020\|EPI_ISL_531501\|20 | ···················· | ···················· | ···················· | ···················· | ···················· | ···················· |
| H | 5671 | hCoV-19/Northern Ireland/QEUH-963D74/2020\|EPI_ISL_ | ···················· | ···················· | ···················· | ···················· | ···················· | ···················· |
| H | 5672 | hCoV-19/England/QEUH-9628DE/2020\|EPI_ISL_531503\|20 | ···················· | ···················· | ···················· | ···················· | ···················· | ···················· |
| H | 5673 | hCoV-19/Scotland/QEUH-96A548/2020\|EPI_ISL_531504\|2 | ···················· | ···················· | ···················· | ···················· | ···················· | ···················· |
| H | 5674 | hCoV-19/England/QEUH-96A7F7/2020\|EPI_ISL_531505\|20 | ···················· | ···················· | ···················· | ···················· | ···················· | ···················· |
| H | 5675 | hCoV-19/England/QEUH-96A6FA/2020\|EPI_ISL_531507\|20 | ···················· | ···················· | ···················· | ···················· | ···················· | ···················· |
| H | 5676 | hCoV-19/England/QEUH-8830EF/2020\|EPI_ISL_532787\|20 | ···················· | ···················· | ···················· | ···················· | ···················· | ···················· |
| H | 5677 | hCoV-19/Wales/QEUH-8AD2BC/2020\|EPI_ISL_532788\|2020 | ···················· | ···················· | ···················· | ···················· | ···················· | ···················· |
| H | 5678 | hCoV-19/England/QEUH-887160/2020\|EPI_ISL_532790\|20 | ···················· | ···················· | ···················· | ···················· | ···················· | ···················· |
| H | 5679 | hCoV-19/Scotland/QEUH-87E8EF/2020\|EPI_ISL_532791\|2 | ···················· | ···················· | ···················· | ···················· | ···················· | ···················· |
| H | 5680 | hCoV-19/England/QEUH-89A434/2020\|EPI_ISL_532792\|20 | ···················· | ···················· | ···················· | ···················· | ···················· | ···················· |
| H | 5681 | hCoV-19/England/QEUH-87DA9F/2020\|EPI_ISL_532793\|20 | ···················· | ···················· | ···················· | ···················· | ···················· | ···················· |
| H | 5682 | hCoV-19/England/QEUH-888396/2020\|EPI_ISL_532794\|20 | ···················· | ···················· | ···················· | ···················· | ···················· | ···················· |
| H | 5683 | hCoV-19/England/QEUH-8B3471/2020\|EPI_ISL_532795\|20 | ···················· | ···················· | ···················· | ···················· | ···················· | ···················· |
| H | 5684 | hCoV-19/England/QEUH-8A7D84/2020\|EPI_ISL_532796\|20 | ···················· | ···················· | ···················· | ···················· | ···················· | ···················· |
| H | 5685 | hCoV-19/England/QEUH-8A02AA/2020\|EPI_ISL_532797\|20 | ···················· | ···················· | ···················· | ···················· | ···················· | ···················· |
| H | 5686 | hCoV-19/England/QEUH-8A8EEA/2020\|EPI_ISL_532798\|20 | ···················· | ···················· | ···················· | ···················· | ···················· | ···················· |
| H | 5687 | hCoV-19/England/QEUH-8970BB/2020\|EPI_ISL_532799\|20 | ···················· | ···················· | ···················· | ···················· | ···················· | ···················· |
| H | 5688 | hCoV-19/England/QEUH-886BEB/2020\|EPI_ISL_532820\|20 | ···················· | ···················· | ···················· | ···················· | ···················· | ···················· |

| | | | | | | | | |
|---|---|---|---|---|---|---|---|---|
| H | 5689 hCoV-19/England/QEUH-88DA9C/2020\|EPI_ISL_532822\|20 | ···················· | ···················· | ····················· | ···················· | ···················· | ···················· | |
| H | 5690 hCoV-19/England/QEUH-88D3CE/2020\|EPI_ISL_532823\|20 | ···················· | ·A·················· | ····················· | ···················· | ···················· | ···················· | |
| H | 5691 hCoV-19/England/QEUH-896B33/2020\|EPI_ISL_532825\|20 | ···················· | ···················· | ····················· | ···················· | ···················· | ···················· | |
| H | 5692 hCoV-19/Northern Ireland/QEUH-88AEFD/2020\|EPI_ISL_ | ···················· | ···················· | ····················· | ···················· | ···················· | ···················· | |
| H | 5693 hCoV-19/Northern Ireland/QEUH-87CACD/2020\|EPI_ISL_ | ···················· | ···················· | ····················· | ···················· | ···················· | ···················· | |
| H | 5694 hCoV-19/England/QEUH-87F408/2020\|EPI_ISL_532828\|20 | ···················· | ·A·················· | ····················· | ···················· | ···················· | ···················· | |
| H | 5695 hCoV-19/England/QEUH-89A66B/2020\|EPI_ISL_532829\|20 | ···················· | ···················· | ····················· | ···················· | ···················· | ···················· | |
| H | 5696 hCoV-19/Northern Ireland/QEUH-889F82/2020\|EPI_ISL_ | ···················· | ···················· | ····················· | ···················· | ···················· | ···················· | |
| H | 5697 hCoV-19/England/QEUH-8AE94D/2020\|EPI_ISL_532831\|20 | ···················· | ···················· | ····················· | ···················· | ···················· | ···················· | |
| H | 5698 hCoV-19/England/QEUH-893E5B/2020\|EPI_ISL_532832\|20 | ···················· | ···················· | ····················· | ···················· | ···················· | ···················· | |
| H | 5699 hCoV-19/Northern Ireland/QEUH-8A88EC/2020\|EPI_ISL_ | ···················· | ···················· | ····················· | ···················· | ···················· | ···················· | |
| H | 5700 hCoV-19/England/QEUH-89E315/2020\|EPI_ISL_532834\|20 | ···················· | ···················· | ····················· | ···················· | ···················· | ···················· | |
| H | 5701 hCoV-19/England/QEUH-7EDA7D/2020\|EPI_ISL_532836\|20 | ···················· | ···················· | ····················· | ···················· | ···················· | ···················· | |
| H | 5702 hCoV-19/Scotland/QEUH-7C5AB7/2020\|EPI_ISL_532838\|2 | ···················· | ···················· | ····················· | ···················· | ···················· | ···················· | |
| H | 5703 hCoV-19/England/OXON-B4CCB/2020\|EPI_ISL_535005\|202 | ···················· | ···················· | ····················· | ···················· | ···················· | ···················· | |
| H | 5704 hCoV-19/England/OXON-B4CE9/2020\|EPI_ISL_535006\|202 | ···················· | ···················· | ····················· | ···················· | ···················· | ···················· | |
| H | 5705 hCoV-19/England/OXON-B4D04/2020\|EPI_ISL_535007\|202 | ···················· | ···················· | ····················· | ···················· | ···················· | ···················· | |
| H | 5706 hCoV-19/England/OXON-B4D13/2020\|EPI_ISL_535008\|202 | ···················· | ···················· | ····················· | ···················· | ···················· | ···················· | |
| H | 5707 hCoV-19/England/OXON-B4D31/2020\|EPI_ISL_535009\|202 | ···················· | ···················· | ····················· | ···················· | ···················· | ···················· | |
| H | 5708 hCoV-19/England/OXON-B4D40/2020\|EPI_ISL_535010\|202 | ···················· | ···················· | ····················· | ···················· | ···················· | ···················· | |
| H | 5709 hCoV-19/England/OXON-B4D5F/2020\|EPI_ISL_535011\|202 | ···················· | ···················· | ····················· | ···················· | ···················· | ···················· | |
| H | 5710 hCoV-19/England/OXON-B4D6E/2020\|EPI_ISL_535012\|202 | ···················· | ···················· | ····················· | ···················· | ···················· | ···················· | |
| H | 5711 hCoV-19/England/OXON-B4D7D/2020\|EPI_ISL_535013\|202 | ···················· | ···················· | ····················· | ···················· | ···················· | ···················· | |
| H | 5712 hCoV-19/England/OXON-B4D8C/2020\|EPI_ISL_535014\|202 | ···················· | ···················· | ····················· | ···················· | ···················· | ···················· | |
| H | 5713 hCoV-19/England/OXON-B4DAA/2020\|EPI_ISL_535015\|202 | ···················· | ···················· | ····················· | ···················· | ···················· | ···················· | |
| H | 5714 hCoV-19/England/OXON-B4DE6/2020\|EPI_ISL_535017\|202 | ···················· | ···················· | ····················· | ···················· | ···················· | ···················· | |
| H | 5715 hCoV-19/England/OXON-B4E01/2020\|EPI_ISL_535018\|202 | ···················· | ···················· | ····················· | ···················· | ···················· | ···················· | |
| H | 5716 hCoV-19/England/OXON-B4E2F/2020\|EPI_ISL_535019\|202 | ···················· | ···················· | ····················· | ···················· | ···················· | ···················· | |
| H | 5717 hCoV-19/Canada/QC-LSPQ-L00242858/2020\|EPI_ISL_5363 | ···················· | ···················· | ····················· | ···················· | ···················· | ···················· | |
| H | 5718 hCoV-19/Canada/QC-LSPQ-L00242867/2020\|EPI_ISL_5363 | ···················· | ···················· | ····················· | ···················· | ···················· | ···················· | |
| H | 5719 hCoV-19/Canada/QC-LSPQ-L00242878/2020\|EPI_ISL_5363 | ···················· | ···················· | ····················· | ···················· | ···················· | ···················· | |
| H | 5720 hCoV-19/Canada/QC-LSPQ-L00242891/2020\|EPI_ISL_5363 | ···················· | ···················· | ····················· | ···················· | ···················· | ···················· | |
| H | 5721 hCoV-19/Canada/QC-LSPQ-L00242900/2020\|EPI_ISL_5363 | ···················· | ···················· | ····················· | ···················· | ···················· | ···················· | |
| H | 5722 hCoV-19/Canada/QC-LSPQ-L00242918/2020\|EPI_ISL_5363 | ···················· | ···················· | ····················· | ···················· | ···················· | ···················· | |
| H | 5723 hCoV-19/Canada/QC-LSPQ-L00243024/2020\|EPI_ISL_5363 | ···················· | ···················· | ····················· | ···················· | ···················· | ···················· | |
| H | 5724 hCoV-19/Canada/QC-LSPQ-L00243054/2020\|EPI_ISL_5363 | ···················· | ···················· | ····················· | ···················· | ···················· | ···················· | |
| H | 5725 hCoV-19/Canada/QC-LSPQ-L00243079/2020\|EPI_ISL_5363 | ···················· | ···················· | ····················· | ···················· | ···················· | ···················· | |
| H | 5726 hCoV-19/Canada/QC-LSPQ-L00243118/2020\|EPI_ISL_5363 | ···················· | ···················· | ····················· | ···················· | ···················· | ···················· | |
| H | 5728 hCoV-19/England/LEED-2AA3C1/2020\|EPI_ISL_538900\|20 | ···················· | ···················· | ····················· | ···················· | ···················· | ···················· | |
| H | 5729 hCoV-19/England/LEED-2AA613/2020\|EPI_ISL_538901\|20 | ···················· | ···················· | ····················· | ···················· | ···················· | ···················· | |
| H | 5730 hCoV-19/England/LEED-2AA2A6/2020\|EPI_ISL_538902\|20 | ···················· | ···················· | ····················· | ···················· | ···················· | ···················· | |
| H | 5731 hCoV-19/England/LEED-2AA200/2020\|EPI_ISL_538903\|20 | ···················· | ·A·················· | ····················· | ···················· | ···················· | ···················· | |
| H | 5732 hCoV-19/England/LEED-2AA516/2020\|EPI_ISL_538904\|20 | ···················· | ···················· | ····················· | ···················· | ···················· | ···················· | |
| H | 5733 hCoV-19/England/LEED-2AA640/2020\|EPI_ISL_538905\|20 | ···················· | ···················· | ····················· | ···················· | ···················· | ···················· | |
| H | 5734 hCoV-19/England/LEED-2AA2B5/2020\|EPI_ISL_538906\|20 | ···················· | ···················· | ····················· | ···················· | ···················· | ···················· | |
| H | 5735 hCoV-19/England/LEED-2A9B06/2020\|EPI_ISL_538907\|20 | ···················· | ···················· | ····················· | ···················· | ···················· | ···················· | |
| H | 5736 hCoV-19/England/LEED-2AA358/2020\|EPI_ISL_538908\|20 | ···················· | ···················A | ····················· | ···················· | ···················· | ···················· | 1 |
| H | 5737 hCoV-19/England/LEED-2A98B4/2020\|EPI_ISL_538909\|20 | ···················· | ···················A | ····················· | ···················· | ···················· | ···················· | 1 |
| H | 5738 hCoV-19/England/LEED-2A98D2/2020\|EPI_ISL_538910\|20 | ···················· | ···················· | ····················· | ···················· | ···················· | ···················· | |
| H | 5739 hCoV-19/England/LEED-2A98A5/2020\|EPI_ISL_538911\|20 | ···················· | ···················· | ····················· | ···················· | ···················· | ···················· | |
| H | 5740 hCoV-19/England/LEED-2A9B42/2020\|EPI_ISL_538912\|20 | ···················· | ·A·················· | ····················· | ···················· | ···················· | ···················· | |
| H | 5741 hCoV-19/England/LEED-2AA112/2020\|EPI_ISL_538913\|20 | ···················· | ···················· | ····················· | ···················· | ···················· | ···················· | |
| H | 5742 hCoV-19/England/LEED-2A990C/2020\|EPI_ISL_538914\|20 | ···················· | ···················· | ····················· | ···················· | ···················· | ···················· | |
| H | 5743 hCoV-19/England/LEED-2AA33A/2020\|EPI_ISL_538915\|20 | ···················· | ···················· | ····················· | ···················· | ···················· | ···················· | |
| H | 5744 hCoV-19/England/LEED-2A98E1/2020\|EPI_ISL_538916\|20 | ···················· | ···················· | ····················· | ···················· | ···················· | ···················· | |
| H | 5745 hCoV-19/England/LEED-2A9869/2020\|EPI_ISL_538918\|20 | ···················· | ···················· | ····················· | ···················· | ···················· | ···················· | |
| H | 5746 hCoV-19/England/LEED-2A99FD/2020\|EPI_ISL_538919\|20 | ···················· | ···················· | ····················· | ···················· | ···················· | ···················· | |
| H | 5747 hCoV-19/England/LEED-2A9ABE/2020\|EPI_ISL_538920\|20 | ···················· | ···················· | ····················· | ···················· | ···················· | ···················· | |
| H | 5748 hCoV-19/England/LEED-2AA07F/2020\|EPI_ISL_538921\|20 | ···················· | ·A·················· | ····················· | ···················· | ···················· | ···················· | |
| H | 5749 hCoV-19/England/LEED-2A9B15/2020\|EPI_ISL_538922\|20 | ···················· | ···················· | ····················· | ···················· | ···················· | ···················· | |
| H | 5750 hCoV-19/England/LEED-2A9C8B/2020\|EPI_ISL_538923\|20 | ···················· | ···················· | ····················· | ···················· | ···················· | ···················· | |
| H | 5751 hCoV-19/England/LEED-2A9AEB/2020\|EPI_ISL_538924\|20 | ···················· | ···················· | ····················· | ···················· | ···················· | ···················· | |
| H | 5752 hCoV-19/England/LEED-2A83E1/2020\|EPI_ISL_538995\|20 | ···················· | ···················A | ····················· | ···················· | ···················· | ···················· | 1 |
| H | 5753 hCoV-19/England/LEED-2A83B4/2020\|EPI_ISL_538996\|20 | ···················· | ···················· | ····················· | ···················· | ···················· | ···················· | |
| H | 5754 hCoV-19/Italy/LOM-UniMI-L106/2020\|EPI_ISL_542104\|2 | ···················· | ···················· | ····················· | ···················· | ···················· | ···················· | |
| H | 5755 hCoV-19/Italy/LOM-UniMI-L107/2020\|EPI_ISL_542105\|2 | ···················· | ···················· | ····················· | ···················· | ···················· | ···················· | |
| H | 5756 hCoV-19/Italy/LOM-UniMI-L108/2020\|EPI_ISL_542106\|2 | ···················· | ···················· | ····················· | ···················· | ···················· | ···················· | |
| H | 5757 hCoV-19/Italy/LOM-UniMI-L109/2020\|EPI_ISL_542107\|2 | ···················· | ···················· | ····················· | ···················· | ···················· | ···················· | |
| H | 5758 hCoV-19/Italy/LOM-UniMI-L111/2020\|EPI_ISL_542108\|2 | ···················· | ···················· | ····················· | ···················· | ···················· | ···················· | |
| H | 5759 hCoV-19/Italy/LOM-UniMI-L112/2020\|EPI_ISL_542109\|2 | ···················· | ···················· | ····················· | ···················· | ···················· | ···················· | |
| H | 5760 hCoV-19/Italy/LOM-UniMI-L113/2020\|EPI_ISL_542110\|2 | ···················· | ···················· | ····················· | ···················· | ···················· | ···················· | |
| H | 5761 hCoV-19/Italy/LOM-UniMI-L115/2020\|EPI_ISL_542111\|2 | ···················· | ···················· | ····················· | ···················· | ···················· | ···················· | |
| H | 5762 hCoV-19/Italy/LOM-UniMI-L116/2020\|EPI_ISL_542112\|2 | ···················· | ···················· | ····················· | ···················· | ···················· | ···················· | |
| H | 5763 hCoV-19/Italy/LOM-UniMI-L117/2020\|EPI_ISL_542113\|2 | ···················· | ···················· | ····················· | ···················· | ···················· | ···················· | |
| H | 5764 hCoV-19/Italy/LOM-UniMI-L118/2020\|EPI_ISL_542114\|2 | ···················· | ···················· | ····················· | ···················· | ···················· | ···················· | |
| H | 5765 hCoV-19/Italy/LOM-UniMI-L119/2020\|EPI_ISL_542115\|2 | ···················· | ···················· | ····················· | ···················· | ···················· | ···················· | |
| H | 5766 hCoV-19/Italy/LOM-UniMI-L120/2020\|EPI_ISL_542116\|2 | ···················· | ···················· | ····················· | ···················· | ···················· | ···················· | |
| H | 5767 hCoV-19/Italy/LOM-UniMI-L121/2020\|EPI_ISL_542117\|2 | ···················· | ···················· | ····················· | ···················· | ···················· | ···················· | |
| H | 5768 hCoV-19/Italy/LOM-UniMI-L122/2020\|EPI_ISL_542118\|2 | ···················· | ···················· | ····················· | ···················· | ···················· | ···················· | |
| H | 5769 hCoV-19/Italy/LOM-UniMI-L123/2020\|EPI_ISL_542119\|2 | ···················· | ···················· | ····················· | ···················· | ···················· | ···················· | |

| | | | | | | | | |
|---|---|---|---|---|---|---|---|---|
| H | 5770 | hCoV-19/Italy/LOM-UniMI-L124/2020\|EPI_ISL_542120\|2 | ···················· | ···················· | ···················· | ···················· | ···················· | ···················· |
| H | 5771 | hCoV-19/Italy/LOM-UniMI-L125/2020\|EPI_ISL_542121\|2 | ···T················ | ···················· | ···················· | ···················· | ···················· | ···················· |
| H | 5772 | hCoV-19/Italy/LOM-UniMI-L126/2020\|EPI_ISL_542122\|2 | ···················· | ···················· | ···················· | ···················· | ···················· | ···················· |
| H | 5773 | hCoV-19/Italy/LOM-UniMI-L127/2020\|EPI_ISL_542123\|2 | ···················· | ···················· | ···················· | ···················· | ···················· | ···················· |
| H | 5774 | hCoV-19/Italy/LOM-UniMI-L128/2020\|EPI_ISL_542124\|2 | ···················· | ···················· | ···················· | ···················· | ···················· | ···················· |
| H | 5775 | hCoV-19/Italy/LOM-UniMI-L129/2020\|EPI_ISL_542125\|2 | ···················· | ···················· | ···················· | ···················· | ···················· | ···················· |
| H | 5776 | hCoV-19/Italy/LOM-UniMI-L130/2020\|EPI_ISL_542126\|2 | ···················· | ···················· | ···················· | ···················· | ···················· | ···················· |
| H | 5777 | hCoV-19/Italy/LOM-UniMI-L131/2020\|EPI_ISL_542127\|2 | ···················· | ···················· | ···················· | ···················· | ···················· | ···················· |
| H | 5778 | hCoV-19/Italy/LOM-UniMI-L132/2020\|EPI_ISL_542128\|2 | ···················· | ···················· | ···················· | ···················· | ···················· | ···················· |
| H | 5779 | hCoV-19/Italy/LOM-UniMI-L133/2020\|EPI_ISL_542129\|2 | ···················· | ···················· | ···················· | ···················· | ···················· | ···················· |
| H | 5780 | hCoV-19/Italy/LOM-UniMI-L134/2020\|EPI_ISL_542130\|2 | ···················· | ···················· | ···················· | ···················· | ···················· | ···················· |
| H | 5781 | hCoV-19/Italy/LOM-UniMI-L135/2020\|EPI_ISL_542131\|2 | ···················· | ···················· | ···················· | ···················· | ···················· | ···················· |
| H | 5782 | hCoV-19/Italy/LOM-UniMI-L136/2020\|EPI_ISL_542132\|2 | ···················· | ···················· | ···················· | ···················· | ···················· | ···················· |
| H | 5783 | hCoV-19/Italy/LOM-UniMI-L137/2020\|EPI_ISL_542133\|2 | ···················· | ···················· | ···················· | ···················· | ···················· | ···················· |
| H | 5784 | hCoV-19/Italy/LOM-UniMI-L138/2020\|EPI_ISL_542134\|2 | ···················· | ···················· | ···················· | ···················· | ···················· | ···················· |
| H | 5785 | hCoV-19/Italy/LOM-UniMI-L139/2020\|EPI_ISL_542135\|2 | ···················· | ···················· | ···················· | ···················· | ···················· | ···················· |
| H | 5786 | hCoV-19/Italy/LOM-UniMI-L140/2020\|EPI_ISL_542136\|2 | ···················· | ···················· | ···················· | ···················· | ···················· | ···················· |
| H | 5787 | hCoV-19/Italy/LOM-UniMI-L141/2020\|EPI_ISL_542137\|2 | ···················· | ···················· | ···················· | ···················· | ···················· | ···················· |
| H | 5788 | hCoV-19/Italy/LOM-UniMI-L142/2020\|EPI_ISL_542138\|2 | ···················· | ···················· | ···················· | ···················· | ···················· | ···················· |
| H | 5789 | hCoV-19/Italy/LOM-UniMI-L144/2020\|EPI_ISL_542139\|2 | ···················· | ···················· | ···················· | ···················· | ···················· | ···················· |
| H | 5790 | hCoV-19/Italy/LOM-UniMI-L145/2020\|EPI_ISL_542140\|2 | ···················· | ···················· | ···················· | ···················· | ···················· | ···················· |
| H | 5791 | hCoV-19/Italy/LOM-UniMI-L146/2020\|EPI_ISL_542141\|2 | ···················· | ···················· | ···················· | ···················· | ···················· | ···················· |
| H | 5792 | hCoV-19/Italy/LOM-UniMI-L147/2020\|EPI_ISL_542142\|2 | ···················· | ···················· | ···················· | ···················· | ···················· | ···················· |
| H | 5793 | hCoV-19/Italy/LOM-UniMI-L148/2020\|EPI_ISL_542143\|2 | ···················· | ···················· | ···················· | ···················· | ···················· | ···················· |
| H | 5794 | hCoV-19/Italy/LOM-UniMI-L149/2020\|EPI_ISL_542144\|2 | ···················· | ···················· | ···················· | ···················· | ···················· | ···················· |
| H | 5795 | hCoV-19/Italy/LOM-UniMI-L150/2020\|EPI_ISL_542145\|2 | ···················· | ···················· | ···················· | ···················· | ···················· | ···················· |
| H | 5796 | hCoV-19/Italy/LOM-UniMI-L151/2020\|EPI_ISL_542146\|2 | ···················· | ···················· | ···················· | ···················· | ···················· | ···················· |
| H | 5797 | hCoV-19/Italy/LOM-UniMI-L152/2020\|EPI_ISL_542147\|2 | ···················· | ···················· | ···················· | ···················· | ···················· | ···················· |
| H | 5798 | hCoV-19/Italy/LOM-UniMI-L153/2020\|EPI_ISL_542148\|2 | ···················· | ···················· | ···················· | ···················· | ···················· | ···················· |
| H | 5799 | hCoV-19/Italy/LOM-UniMI-L154/2020\|EPI_ISL_542149\|2 | ···················· | ···················· | ···················· | ···················· | ···················· | ···················· |
| H | 5800 | hCoV-19/Italy/LOM-UniMI-L155/2020\|EPI_ISL_542150\|2 | ···················· | ···················· | ···················· | ···················· | ···················· | ···················· |
| H | 5801 | hCoV-19/Italy/LOM-UniMI-L156/2020\|EPI_ISL_542151\|2 | ···················· | ···················· | ···················· | ···················· | ···················· | ···················· |
| H | 5802 | hCoV-19/Italy/LOM-UniMI-L157/2020\|EPI_ISL_542152\|2 | ···················· | ···················· | ···················· | ···················· | ···················· | ···················· |
| H | 5803 | hCoV-19/Italy/LOM-UniMI-L158/2020\|EPI_ISL_542153\|2 | ···················· | ···················· | ···················· | ···················· | ···················· | ···················· |
| H | 5804 | hCoV-19/Italy/LOM-UniMI-L159/2020\|EPI_ISL_542154\|2 | ···················· | ···················· | ···················· | ···················· | ···················· | ···················· |
| H | 5805 | hCoV-19/Italy/LOM-UniMI-L160/2020\|EPI_ISL_542155\|2 | ···················· | ···················· | ···················· | ···················· | ···················· | ···················· |
| H | 5806 | hCoV-19/Italy/LOM-UniMI-L162/2020\|EPI_ISL_542156\|2 | ···················· | ···················· | ···················· | ···················· | ···················· | ···················· |
| H | 5807 | hCoV-19/Italy/LOM-UniMI-L163/2020\|EPI_ISL_542157\|2 | ···················· | ···················· | ···················· | ···················· | ···················· | ···················· |
| H | 5808 | hCoV-19/Italy/LOM-UniMI-L164/2020\|EPI_ISL_542158\|2 | ···················· | ···················· | ···················· | ···················· | ···················· | ···················· |
| H | 5809 | hCoV-19/Italy/LOM-UniMI-L165/2020\|EPI_ISL_542159\|2 | ···················· | ···················· | ···················· | ···················· | ···················· | ···················· |
| H | 5810 | hCoV-19/Italy/LOM-UniMI-L166/2020\|EPI_ISL_542160\|2 | ···················· | ···················· | ···················· | ···················· | ···················· | ···················· |
| H | 5811 | hCoV-19/Italy/LOM-UniMI-L167/2020\|EPI_ISL_542161\|2 | ···················· | ···················· | ···················· | ···················· | ···················· | ···················· |
| H | 5812 | hCoV-19/Italy/LOM-UniMI-L168/2020\|EPI_ISL_542162\|2 | ···················· | ···················· | ···················· | ···················· | ···················· | ···················· |
| H | 5813 | hCoV-19/Italy/LOM-UniMI-L169/2020\|EPI_ISL_542163\|2 | ···················· | ···················· | ···················· | ···················· | ···················· | ···················· |
| H | 5814 | hCoV-19/Italy/LOM-UniMI-L170/2020\|EPI_ISL_542164\|2 | ···················· | ···················· | ···················· | ···················· | ···················· | ···················· |
| H | 5815 | hCoV-19/Italy/LOM-UniMI-L171/2020\|EPI_ISL_542165\|2 | ···················· | ···················· | ···················· | ···················· | ···················· | ···················· |
| H | 5816 | hCoV-19/Italy/LOM-UniMI-L172/2020\|EPI_ISL_542166\|2 | ···················· | ···················· | ···················· | ···················· | ···················· | ···················· |
| H | 5817 | hCoV-19/Italy/LOM-UniMI-L173/2020\|EPI_ISL_542167\|2 | ···················· | ···················· | ···················· | ···················· | ···················· | ···················· |
| H | 5818 | hCoV-19/Italy/LOM-UniMI-L174/2020\|EPI_ISL_542168\|2 | ···················· | ···················· | ···················· | ···················· | ···················· | ···················· |
| H | 5819 | hCoV-19/Italy/LOM-UniMI-L175/2020\|EPI_ISL_542169\|2 | ···················· | ···················· | ···················· | ···················· | ···················· | ···················· |
| H | 5820 | hCoV-19/Italy/LOM-UniMI-L176/2020\|EPI_ISL_542170\|2 | ···················· | ···················· | ···················· | ···················· | ···················· | ···················· |
| H | 5821 | hCoV-19/Italy/LOM-UniMI-L177/2020\|EPI_ISL_542171\|2 | ···················· | ···················· | ···················· | ···················· | ···················· | ···················· |
| H | 5822 | hCoV-19/Italy/LOM-UniMI-L178/2020\|EPI_ISL_542172\|2 | ···················· | ···················· | ···················· | ···················· | ···················· | ···················· |
| H | 5823 | hCoV-19/Italy/LOM-UniMI-L182/2020\|EPI_ISL_542173\|2 | ···················· | ···················· | ···················· | ···················· | ···················· | ···················· |
| H | 5824 | hCoV-19/Italy/LOM-UniMI-L183/2020\|EPI_ISL_542174\|2 | ···················· | ···················· | ···················· | ···················· | ···················· | ···················· |
| H | 5825 | hCoV-19/Italy/LOM-UniMI-L184/2020\|EPI_ISL_542175\|2 | ···················· | ···················· | ···················· | ···················· | ···················· | ···················· |
| H | 5826 | hCoV-19/Italy/LOM-UniMI-L185/2020\|EPI_ISL_542176\|2 | ···················· | ···················· | ···················· | ···················· | ···················· | ···················· |
| H | 5827 | hCoV-19/Italy/LOM-UniMI-L186/2020\|EPI_ISL_542177\|2 | ···················· | ···················· | ···················· | ···················· | ···················· | ···················· |
| H | 5828 | hCoV-19/Italy/LOM-UniMI-L187/2020\|EPI_ISL_542178\|2 | ···················· | ···················· | ···················· | ···················· | ···················· | ···················· |
| H | 5829 | hCoV-19/Italy/LOM-UniMI-L189/2020\|EPI_ISL_542179\|2 | ···················· | ···················· | ···················· | ···················· | ···················· | ···················· |
| H | 5830 | hCoV-19/Italy/LOM-UniMI-L190/2020\|EPI_ISL_542180\|2 | ···················· | ···················· | ···················· | ···················· | ···················· | ···················· |
| H | 5831 | hCoV-19/Italy/LOM-UniMI-L191/2020\|EPI_ISL_542181\|2 | ···················· | ···················· | ···················· | ···················· | ···················· | ···················· |
| H | 5832 | hCoV-19/Italy/LOM-UniMI-L192/2020\|EPI_ISL_542182\|2 | ···················· | ···················· | ···················· | ···················· | ···················· | ···················· |
| H | 5833 | hCoV-19/Italy/LOM-UniMI-L193/2020\|EPI_ISL_542183\|2 | ···················· | ···················· | ···················· | ···················· | ···················· | ···················· |
| H | 5834 | hCoV-19/Italy/LOM-UniMI-L194/2020\|EPI_ISL_542184\|2 | ···················· | ···················· | ···················· | ···················· | ···················· | ···················· |
| H | 5835 | hCoV-19/Italy/LOM-UniMI-L195/2020\|EPI_ISL_542185\|2 | ···················· | ···················· | ···················· | ···················· | ···················· | ···················· |
| H | 5836 | hCoV-19/Italy/LOM-UniMI-L196/2020\|EPI_ISL_542186\|2 | ···················· | ···················· | ···················· | ···················· | ···················· | ···················· |
| H | 5837 | hCoV-19/Italy/LOM-UniMI-L197/2020\|EPI_ISL_542187\|2 | ···················· | ···················· | ···················· | ···················· | ···················· | ···················· |
| H | 5838 | hCoV-19/Italy/LOM-UniMI-L198/2020\|EPI_ISL_542188\|2 | ···················· | ···················· | ···················· | ···················· | ···················· | ···················· |
| H | 5839 | hCoV-19/Italy/LOM-UniMI-L199/2020\|EPI_ISL_542189\|2 | ···················· | ···················· | ···················· | ···················· | ···················· | ···················· |
| H | 5840 | hCoV-19/USA/TX-HMH-4204/2020\|EPI_ISL_543435\|2020-0 | ···················· | ···················· | ···················· | ···················· | ···················· | ···················· |
| H | 5842 | hCoV-19/USA/TX-HMH-4206/2020\|EPI_ISL_543437\|2020-0 | ···················· | ···················· | ···················· | ···················· | ···················· | N··················· |
| H | 5843 | hCoV-19/USA/TX-HMH-4207/2020\|EPI_ISL_543438\|2020-0 | ···················· | ···················· | ···················· | ···················· | ···················· | ···················· |
| H | 5844 | hCoV-19/USA/TX-HMH-4208/2020\|EPI_ISL_543439\|2020-0 | ···················· | ···················· | ···················· | ···················· | ···················· | ···················· |
| H | 5845 | hCoV-19/USA/TX-HMH-4209/2020\|EPI_ISL_543440\|2020-0 | ···················· | ···················· | ···················· | ···················· | ···················· | ···················· |
| H | 5846 | hCoV-19/USA/TX-HMH-4210/2020\|EPI_ISL_543441\|2020-0 | ···················· | ···················· | ···················· | ···················· | ···················· | ···················· |
| H | 5847 | hCoV-19/USA/TX-HMH-4212/2020\|EPI_ISL_543442\|2020-0 | ···················· | ···················· | ···················· | ···················· | ···················· | ···················· |
| H | 5848 | hCoV-19/USA/TX-HMH-4213/2020\|EPI_ISL_543443\|2020-0 | ···················· | ···················· | ···················· | ···················· | ···················· | ···················· |
| H | 5855 | hCoV-19/USA/TX-HMH-4242/2020\|EPI_ISL_543455\|2020-0 | ···················· | ···················· | ···················· | ···················· | ···················· | ···················· |
| H | 5856 | hCoV-19/USA/TX-HMH-4243/2020\|EPI_ISL_543456\|2020-0 | ···················· | ···················· | ···················· | ···················· | ···················· | ···················· |

H 5857 hCoV-19/USA/TX-HMH-4245/2020|EPI_ISL_543458|2020-0 ···················· ·A·················· ····················· ···················· ···················· ····················
H 5859 hCoV-19/USA/TX-HMH-4248/2020|EPI_ISL_543460|2020-0 ···················· ···················· ····················· ···················· ···················· ····················
H 5860 hCoV-19/USA/TX-HMH-4249/2020|EPI_ISL_543461|2020-0 ···················· ···················· ····················· ···················· ···················· ····················
H 5861 hCoV-19/USA/TX-HMH-4252/2020|EPI_ISL_543464|2020-0 ···················· ···················· ····················· ···················· ··N···········N····· ··N·················
H 5865 hCoV-19/USA/TX-HMH-4257/2020|EPI_ISL_543469|2020-0 ···················· ·············A······ ····················· ···················· ···················· ····················
H 5866 hCoV-19/USA/TX-HMH-4258/2020|EPI_ISL_543470|2020-0 ···················· ···················· ····················· ···················· ···················· ····················
H 5867 hCoV-19/USA/TX-HMH-4259/2020|EPI_ISL_543471|2020-0 ···················· ···················· ····················· ···················· ···················· ····················
H 5868 hCoV-19/USA/TX-HMH-4261/2020|EPI_ISL_543473|2020-0 ···················· ·A·················· ····················· ···················· ···················· ····················
H 5869 hCoV-19/USA/TX-HMH-4262/2020|EPI_ISL_543474|2020-0 ···················· ···················· ····················· ···················· ···················· ····················
H 5870 hCoV-19/USA/TX-HMH-4263/2020|EPI_ISL_543475|2020-0 ···················· ···················· ····················· ···················· ···················· ····················
H 5872 hCoV-19/USA/TX-HMH-4265/2020|EPI_ISL_543477|2020-0 ···················· ···················· ····················· ···················· ···················· ····················
H 5873 hCoV-19/USA/TX-HMH-4266/2020|EPI_ISL_543478|2020-0 ···················· ···················· ····················· ···················· ···················· ····················
H 5874 hCoV-19/USA/TX-HMH-4267/2020|EPI_ISL_543479|2020-0 ···················· ···················· ····················· ···················· ···················· ····················
H 5878 hCoV-19/USA/TX-HMH-4274/2020|EPI_ISL_543486|2020-0 ···················· ···················· ····················· ···················· ···················· ····················
H 5879 hCoV-19/USA/TX-HMH-4275/2020|EPI_ISL_543487|2020-0 ···················· ···················· ····················· ···················· ···················· ····················
H 5882 hCoV-19/USA/TX-HMH-4278/2020|EPI_ISL_543490|2020-0 ···················· ···················· ····················· ···················· ···················· ····················
H 5883 hCoV-19/USA/TX-HMH-4279/2020|EPI_ISL_543491|2020-0 ···················· ···················· ····················· ···················· ···················· ····················
H 5884 hCoV-19/USA/TX-HMH-4282/2020|EPI_ISL_543493|2020-0 ···················· ·············A······ ····················· ···················· ···················· ····················
H 5885 hCoV-19/USA/TX-HMH-4283/2020|EPI_ISL_543494|2020-0 ···················· ···················· ····················· ···················· ···················· ····················
H 5886 hCoV-19/USA/TX-HMH-4284/2020|EPI_ISL_543495|2020-0 ···················· ···················· ····················· ···················· ···················· ··A·················
H 5887 hCoV-19/USA/TX-HMH-4287/2020|EPI_ISL_543497|2020-0 ···················· ···············N···· ····················· ···················· ···················· ····················
H 5888 hCoV-19/USA/TX-HMH-4288/2020|EPI_ISL_543498|2020-0 ···················· ···················· ····················· ···················· ···················· ····················
H 5889 hCoV-19/Australia/QLD959/2020|EPI_ISL_530225|2020- ···················· ···················· ····················· ···················· ···················· ····················
H 5890 hCoV-19/Australia/QLD963/2020|EPI_ISL_530226|2020- ···················· ···················· ····················· ···················· ···················· ····················
H 5891 hCoV-19/Australia/QLD979/2020|EPI_ISL_530227|2020- ···················· ···················· ····················· ···················· ···················· ····················
H 5892 hCoV-19/Australia/QLD968/2020|EPI_ISL_530228|2020- ···················· ···················· ····················· ···················· ···················· ····················
H 5893 hCoV-19/Australia/QLD978/2020|EPI_ISL_530229|2020- ···················· ···················· ····················· ···················· ···················· ····················
H 5894 hCoV-19/Australia/QLD970/2020|EPI_ISL_530230|2020- ···················· ···················· ····················· ···················· ···················· ····················
H 5895 hCoV-19/Australia/QLD973/2020|EPI_ISL_530231|2020- ···················· ···················· ····················· ···················· ···················· ····················
H 5896 hCoV-19/Australia/QLD967/2020|EPI_ISL_530232|2020- ···················· ···················· ····················· ···················· ···················· ····················
H 5897 hCoV-19/Australia/QLD969/2020|EPI_ISL_530233|2020- ···················· ···················· ····················· ···················· ···················· ····················
H 5898 hCoV-19/Australia/QLD966/2020|EPI_ISL_530234|2020- ···················· ···················· ····················· ···················· ···················· ····················
H 5899 hCoV-19/Australia/QLD971/2020|EPI_ISL_530235|2020- ···················· ···················· ····················· ···················· ···················· ····················
H 5900 hCoV-19/Australia/QLD961/2020|EPI_ISL_530236|2020- ···················· ···················· ····················· ···················· ···················· ····················
H 5901 hCoV-19/Australia/QLD974/2020|EPI_ISL_530237|2020- ···················· ···················· ····················· ···················· ···················· ····················
H 5902 hCoV-19/Australia/QLD964/2020|EPI_ISL_530238|2020- ···················· ···················· ····················· ···················· ···················· ····················
H 5903 hCoV-19/Australia/QLD958/2020|EPI_ISL_530239|2020- ···················· ···················· ····················· ···················· ···················· ····················
H 5904 hCoV-19/Australia/QLD957/2020|EPI_ISL_530240|2020- ···················· ···················· ····················· ···················· ···················· ····················
H 5905 hCoV-19/Australia/QLD962/2020|EPI_ISL_530241|2020- ···················· ···················· ····················· ···················· ···················· ····················
H 5906 hCoV-19/Australia/QLD960/2020|EPI_ISL_530242|2020- ···················· ···················· ····················· ···················· ···················· ····················
H 5907 hCoV-19/Australia/QLD955/2020|EPI_ISL_530243|2020- ···················· ···················· ····················· ···················· ···················· ····················
H 5908 hCoV-19/Australia/QLD965/2020|EPI_ISL_530244|2020- ···················· ···················· ····················· ···················· ···················· ····················
H 5909 hCoV-19/Australia/QLD1003/2020|EPI_ISL_530245|2020 ···················· ···················· ····················· ···················· ···················· ····················
H 5910 hCoV-19/Northern Ireland/QEUH-963020/2020|EPI_ISL_ ···················· ···················· ····················· ···················· ···················· ····················
H 5911 hCoV-19/England/QEUH-963BC5/2020|EPI_ISL_531508|20 ···················· ···················· ····················· ···················· ···················· ····················
H 5912 hCoV-19/England/QEUH-96312D/2020|EPI_ISL_531509|20 ···················· ···················· ····················· ···················· ···················· ····················
H 5913 hCoV-19/England/QEUH-963D65/2020|EPI_ISL_531510|20 ···················· ···················· ····················· ···················· ···················· ····················
H 5914 hCoV-19/England/QEUH-96A5A2/2020|EPI_ISL_531511|20 ···················· ···················· ····················· ···················· ···················· ····················
H 5915 hCoV-19/England/QEUH-962847/2020|EPI_ISL_531512|20 ···················· ···················· ····················· ···················· ···················· ····················
H 5916 hCoV-19/England/QEUH-96A5FD/2020|EPI_ISL_531513|20 ···················· ···················· ····················· ···················· ···················· ····················
H 5917 hCoV-19/England/QEUH-96A42D/2020|EPI_ISL_531514|20 ···················· ···················· ····················· ···················· ···················· ····················
H 5918 hCoV-19/England/QEUH-9639F8/2020|EPI_ISL_531515|20 ···················· ···················· ····················· ···················· ···················· ····················
H 5919 hCoV-19/England/QEUH-963D47/2020|EPI_ISL_531516|20 ···················· ···················· ····················· ···················· ···················· ····················
H 5920 hCoV-19/England/QEUH-96A4C3/2020|EPI_ISL_531517|20 ···················· ···················· ····················· ···················· ···················· ····················
H 5921 hCoV-19/England/QEUH-963CA4/2020|EPI_ISL_531518|20 ···················· ···················· ····················· ···················· ···················· ····················
H 5922 hCoV-19/England/QEUH-963011/2020|EPI_ISL_531519|20 ···················· ···················· ····················· ···················· ···················· ····················
H 5923 hCoV-19/England/QEUH-96A478/2020|EPI_ISL_531520|20 ···················· ···················· ····················· ···················· ···················· ····················
H 5924 hCoV-19/England/QEUH-963CC2/2020|EPI_ISL_531521|20 ···················· ···················· ····················· ···················· ···················· ····················
H 5925 hCoV-19/England/QEUH-963B98/2020|EPI_ISL_531522|20 ···················· ···················· ····················· ···················· ···················· ····················
H 5926 hCoV-19/Scotland/QEUH-963E08/2020|EPI_ISL_531524|2 ···················· ···················· ····················· ···················· ···················· ····················
H 5927 hCoV-19/England/QEUH-96A86D/2020|EPI_ISL_531525|20 ···················· ···················· ····················· ···················· ···················· ····················
H 5928 hCoV-19/England/QEUH-96A5B1/2020|EPI_ISL_531526|20 ···················· ···················· ····················· ···················· ···················· ····················
H 5929 hCoV-19/England/QEUH-96A539/2020|EPI_ISL_531528|20 ···················· ···················· ····················· ···················· ···················· ····················
H 5930 hCoV-19/England/QEUH-96A6AF/2020|EPI_ISL_531529|20 ···················· ···················· ····················· ···················· ···················· ····················
H 5931 hCoV-19/England/QEUH-96A575/2020|EPI_ISL_531530|20 ···················· ···················· ····················· ···················· ···················· ····················
H 5932 hCoV-19/England/QEUH-963B2F/2020|EPI_ISL_531531|20 ···················· ···················· ····················· ···················· ···················· ····················
H 5933 hCoV-19/Northern Ireland/QEUH-96308A/2020|EPI_ISL_ ···················· ···················· ····················· ···················· ···················· ····················
H 5934 hCoV-19/Northern Ireland/QEUH-96A52A/2020|EPI_ISL_ ···················· ···················· ····················· ···················· ···················· ····················
H 5935 hCoV-19/England/QEUH-96A812/2020|EPI_ISL_531534|20 ···················· ·A·················· ····················· ···················· ···················· ····················
H 5936 hCoV-19/England/QEUH-963970/2020|EPI_ISL_531535|20 ···················· ···················· ····················· ···················· ···················· ····················
H 5937 hCoV-19/England/QEUH-96A4A5/2020|EPI_ISL_531537|20 ···················· ·A·················· ····················· ···················· ···················· ····················
H 5938 hCoV-19/England/QEUH-962892/2020|EPI_ISL_531538|20 ···················· ···················· ····················· ···················· ···················· ····················
H 5939 hCoV-19/England/QEUH-963D56/2020|EPI_ISL_531539|20 ···················· ·A·················· ····················· ···················· ···················· ··A·················
H 5940 hCoV-19/England/QEUH-96A733/2020|EPI_ISL_531540|20 ···················· ···················· ····················· ···················· ···················· ····················
H 5941 hCoV-19/England/QEUH-963CB3/2020|EPI_ISL_531541|20 ···················· ···················· ····················· ···················· ···················· ····················
H 5942 hCoV-19/England/QEUH-963E62/2020|EPI_ISL_531542|20 ···················· ···················· ····················· ···················· ···················· ····················
H 5943 hCoV-19/England/QEUH-963D38/2020|EPI_ISL_531543|20 ···················· ···················· ····················· ···················· ···················· ····················
H 5944 hCoV-19/England/QEUH-96A44B/2020|EPI_ISL_531544|20 ···················· ···················· ····················· ···················· ···················· ····················
H 5945 hCoV-19/England/QEUH-963CFF/2020|EPI_ISL_531545|20 ···················· ···················· ····················· ···················· ···················· ····················
H 5946 hCoV-19/Scotland/QEUH-96314B/2020|EPI_ISL_531546|2 ···················· ···················· ····················· ···················· ···················· ····················

| | | | | | | | | | |
|---|---|---|---|---|---|---|---|---|---|
| H | 5947 | hCoV-19/Scotland/QEUH-96315A/2020\|EPI_ISL_531548\|2 | ···················· | ···················· | ···················· | ···················· | ···················· | ···················· | |
| H | 5948 | hCoV-19/England/QEUH-963C3B/2020\|EPI_ISL_531550\|20 | ···················· | ···················· | ···················· | ···················· | ···················· | ···················· | |
| H | 5949 | hCoV-19/England/QEUH-96A4B4/2020\|EPI_ISL_531551\|20 | ···················· | ···················· | ···················· | ···················· | ···················· | ···················· | |
| H | 5950 | hCoV-19/England/QEUH-9628B0/2020\|EPI_ISL_531552\|20 | ···················· | ···················· | ···················· | ···················· | ···················· | ···················· | |
| H | 5951 | hCoV-19/England/QEUH-96A5DF/2020\|EPI_ISL_531553\|20 | ···················· | ···················· | ···················· | ···················· | ···················· | ···················· | |
| H | 5952 | hCoV-19/England/QEUH-963B3E/2020\|EPI_ISL_531554\|20 | ···················· | ···················· | ···················· | ···················· | ···················· | ···················· | |
| H | 5953 | hCoV-19/England/QEUH-96A706/2020\|EPI_ISL_531555\|20 | ···················· | ···················· | ···················· | ···················· | ···················· | ···················· | |
| H | 5954 | hCoV-19/Scotland/QEUH-9639BC/2020\|EPI_ISL_531557\|2 | ···················· | ···················· | ···················· | ···················· | ···················· | ···················· | |
| H | 5955 | hCoV-19/England/QEUH-96A760/2020\|EPI_ISL_531558\|20 | ···················· | ···················· | ···················· | ···················· | ···················· | ···················· | |
| H | 5956 | hCoV-19/England/QEUH-96A5EE/2020\|EPI_ISL_531559\|20 | ···················· | ···················· | ···················· | ···················· | ···················· | ···················· | |
| H | 5957 | hCoV-19/England/QEUH-9628CF/2020\|EPI_ISL_531560\|20 | ···················· | ···················· | ···················· | ···················· | ···················· | ···················· | |
| H | 5958 | hCoV-19/England/QEUH-96A724/2020\|EPI_ISL_531561\|20 | ···················· | ···················· | ···················· | ···················· | ··················C· | ···················· | 1 |
| H | 5959 | hCoV-19/England/QEUH-963BF2/2020\|EPI_ISL_531562\|20 | ···················· | ···················· | ···················· | ···················· | ···················· | ···················· | |
| H | 5960 | hCoV-19/Scotland/QEUH-963E17/2020\|EPI_ISL_531563\|2 | ···················· | ···················· | ···················· | ···················· | ···················· | ···················· | |
| H | 5961 | hCoV-19/Scotland/QEUH-963BE3/2020\|EPI_ISL_531564\|2 | ···················· | ···················· | ···················· | ···················· | ···················· | ···················· | |
| H | 5962 | hCoV-19/Scotland/QEUH-96A663/2020\|EPI_ISL_531565\|2 | ···················· | ···················· | ···················· | ···················· | ···················· | ···················· | |
| H | 5963 | hCoV-19/Northern Ireland/QEUH-9639DA/2020\|EPI_ISL_ | ···················· | ···················· | ···················· | ···················· | ···················· | ···················· | |
| H | 5964 | hCoV-19/Scotland/QEUH-963A22/2020\|EPI_ISL_531569\|2 | ···················· | ···················· | ···················· | ···················· | ···················· | ···················· | |
| H | 5965 | hCoV-19/Scotland/QEUH-96A672/2020\|EPI_ISL_531570\|2 | ···················· | ···················· | ···················· | ···················· | ···················· | ···················· | |
| H | 5966 | hCoV-19/England/QEUH-96A557/2020\|EPI_ISL_531571\|20 | ···················· | ···················· | ···················· | ···················· | ···················· | ···················· | |
| H | 5967 | hCoV-19/England/QEUH-96A51B/2020\|EPI_ISL_531573\|20 | ···················· | ···················· | ···················· | ···················· | ···················· | ···················· | |
| H | 5968 | hCoV-19/England/QEUH-96A77F/2020\|EPI_ISL_531574\|20 | ···················· | ···················· | ···················· | ···················· | ···················· | ···················· | |
| H | 5969 | hCoV-19/Scotland/QEUH-96303F/2020\|EPI_ISL_531575\|2 | ···················· | ···················· | ···················· | ···················· | ···················· | ···················· | |
| H | 5970 | hCoV-19/England/QEUH-96A6BE/2020\|EPI_ISL_531576\|20 | ···················· | ···················· | ···················· | ···················· | ···················· | ···················· | |
| H | 5971 | hCoV-19/England/QEUH-7CBF67/2020\|EPI_ISL_532837\|20 | ···················· | ···················· | ···················· | ···················· | ···················· | ···················· | |
| H | 5972 | hCoV-19/England/QEUH-7F31BB/2020\|EPI_ISL_532840\|20 | ···················· | ···················· | ···················· | ···················· | ···················· | ···················· | |
| H | 5973 | hCoV-19/Northern Ireland/QEUH-7DEE91/2020\|EPI_ISL_ | ···················· | ···················· | ···················· | ···················· | ···················· | ···················· | |
| H | 5974 | hCoV-19/England/QEUH-7B2B32/2020\|EPI_ISL_532845\|20 | ···················· | ···················· | ···················· | ···················· | ···················· | ···················· | |
| H | 5975 | hCoV-19/England/QEUH-7F32B8/2020\|EPI_ISL_532847\|20 | ···················· | ···················· | ···················· | ···················· | ···················· | ···················· | |
| H | 5976 | hCoV-19/England/QEUH-7E1E31/2020\|EPI_ISL_532848\|20 | ···················· | ···················· | ···················· | ···················· | ···················· | ···················· | |
| H | 5977 | hCoV-19/England/QEUH-7BBA0F/2020\|EPI_ISL_532849\|20 | ···················· | ···················· | ···················· | ···················· | ···················· | ···················· | |
| H | 5978 | hCoV-19/England/QEUH-7C8CEB/2020\|EPI_ISL_532853\|20 | ···················· | ···················· | ···················· | ···················· | ···················· | ···················· | |
| H | 5979 | hCoV-19/Northern Ireland/QEUH-7C69D7/2020\|EPI_ISL_ | ···················· | ···················· | ···················· | ···················· | ···················· | ···················· | |
| H | 5980 | hCoV-19/England/QEUH-7D2923/2020\|EPI_ISL_532855\|20 | ···················· | ···················· | ···················· | ···················· | ···················· | ···················· | |
| H | 5981 | hCoV-19/England/QEUH-7AE8E7/2020\|EPI_ISL_532856\|20 | ···················· | ·A·················· | ···················· | ···················· | ···················· | ···················· | |
| H | 5982 | hCoV-19/England/QEUH-7AED33/2020\|EPI_ISL_532860\|20 | ···················· | ···················· | ···················· | ···················· | ···················· | ···················· | |
| H | 5983 | hCoV-19/England/QEUH-7E5E6A/2020\|EPI_ISL_532861\|20 | ···················· | ···················· | ···················· | ···················· | ···················· | ···················· | |
| H | 5984 | hCoV-19/England/QEUH-7EEE43/2020\|EPI_ISL_532863\|20 | ···················· | ···················· | ···················· | ···················· | ···················· | ···················· | |
| H | 5985 | hCoV-19/Scotland/QEUH-7FD48B/2020\|EPI_ISL_532865\|2 | ···················· | ···················· | ···················· | ···················· | ···················· | ···················· | |
| H | 5986 | hCoV-19/England/QEUH-7E22BC/2020\|EPI_ISL_532866\|20 | ···················· | ·A·················· | ···················· | ···················· | ···················· | ···················· | |
| H | 5987 | hCoV-19/England/QEUH-7ED846/2020\|EPI_ISL_532867\|20 | ···················· | ···················· | ···················· | ···················· | ···················· | ···················· | |
| H | 5988 | hCoV-19/England/QEUH-7EE71B/2020\|EPI_ISL_532868\|20 | ···················· | ···················· | ···················· | ···················· | ···················· | ···················· | |
| H | 5989 | hCoV-19/England/QEUH-7FE226/2020\|EPI_ISL_532869\|20 | ···················· | ···················· | ···················· | ···················· | ···················· | ···················· | |
| H | 5990 | hCoV-19/England/QEUH-7B20BB/2020\|EPI_ISL_532870\|20 | ···················· | ···················· | ···················· | ···················· | ···················· | ···················· | |
| H | 5991 | hCoV-19/England/QEUH-7FA916/2020\|EPI_ISL_532871\|20 | ···················· | ···················· | ···················· | ···················· | ···················· | ···················· | |
| H | 5992 | hCoV-19/England/QEUH-7F7120/2020\|EPI_ISL_532872\|20 | ···················· | ···················· | ···················· | ···················· | ···················· | ···················· | |
| H | 5993 | hCoV-19/Scotland/QEUH-7AC3BB/2020\|EPI_ISL_532874\|2 | ···················· | ···················· | ···················· | ···················· | ···················· | ···················· | |
| H | 5994 | hCoV-19/Scotland/QEUH-7B078B/2020\|EPI_ISL_532876\|2 | ···················· | ···················· | ···················· | ···················· | ···················· | ···················· | |
| H | 5995 | hCoV-19/England/QEUH-7F9C78/2020\|EPI_ISL_532877\|20 | ···················· | ···················· | ···················· | ···················· | ···················· | ···················· | |
| H | 5996 | hCoV-19/Scotland/QEUH-7B57D1/2020\|EPI_ISL_532878\|2 | ···················· | ···················· | ···················· | ···················· | ···················· | ···················· | |
| H | 5997 | hCoV-19/England/QEUH-7D7FB3/2020\|EPI_ISL_532879\|20 | ···················· | ···················· | ···················· | ···················· | ···················· | ···················· | |
| H | 5998 | hCoV-19/England/QEUH-7F000C/2020\|EPI_ISL_532882\|20 | ···················· | ···················· | ···················· | ···················· | ···················· | ···················· | |
| H | 5999 | hCoV-19/England/QEUH-7AF673/2020\|EPI_ISL_532883\|20 | ···················· | ·········A·········· | ···················· | ···················· | ···················· | ···················· | |
| H | 6000 | hCoV-19/England/QEUH-7F9AAB/2020\|EPI_ISL_532884\|20 | ···················· | ···················· | ···················· | ···················· | ···················· | ···················· | |
| H | 6001 | hCoV-19/Scotland/QEUH-7D36D0/2020\|EPI_ISL_532885\|2 | ···················· | ···················· | ···················· | ···················· | ···················· | ···················· | |
| H | 6002 | hCoV-19/England/QEUH-7F2B36/2020\|EPI_ISL_532887\|20 | ···················· | ···················· | ···················· | ···················· | ···················· | ···················· | |
| H | 6003 | hCoV-19/England/QEUH-7C9048/2020\|EPI_ISL_532888\|20 | ···················· | ···················· | ···················· | ···················· | ···················· | ···················· | |
| H | 6004 | hCoV-19/England/QEUH-7EACF2/2020\|EPI_ISL_532889\|20 | ···················· | ···················· | ···················· | ···················· | ···················· | ···················· | |
| H | 6005 | hCoV-19/England/QEUH-7B73CC/2020\|EPI_ISL_532890\|20 | ···················· | ···················· | ···················· | ···················· | ···················· | ···················· | |
| H | 6006 | hCoV-19/Scotland/QEUH-7D35A6/2020\|EPI_ISL_532891\|2 | ···················· | ···················· | ···················· | ···················· | ···················· | ···················· | |
| H | 6007 | hCoV-19/England/QEUH-7DADD4/2020\|EPI_ISL_532892\|20 | ···················· | ···················· | ···················· | ···················· | ···················· | ···················· | |
| H | 6008 | hCoV-19/England/QEUH-7BE170/2020\|EPI_ISL_532893\|20 | ···················· | ···················· | ···················· | ···················· | ···················· | ···················· | |
| H | 6009 | hCoV-19/Scotland/QEUH-7B19B1/2020\|EPI_ISL_532894\|2 | ···················· | ···················· | ···················· | ···················· | ···················· | ···················· | |
| H | 6010 | hCoV-19/England/QEUH-7C016C/2020\|EPI_ISL_532895\|20 | ···················· | ···················· | ···················· | ···················· | ···················· | ···················· | |
| H | 6011 | hCoV-19/Scotland/QEUH-7BC303/2020\|EPI_ISL_532896\|2 | ···················· | ···················· | ···················· | ···················· | ···················· | ···················· | |
| H | 6012 | hCoV-19/Northern Ireland/QEUH-7EC980/2020\|EPI_ISL_ | ···················· | ···················· | ···················· | ···················· | ···················· | ···················· | |
| H | 6013 | hCoV-19/England/QEUH-7CBAFD/2020\|EPI_ISL_532898\|20 | ···················· | ···················· | ···················· | ···················· | ···················· | ···················· | |
| H | 6014 | hCoV-19/England/QEUH-7EB7A5/2020\|EPI_ISL_532899\|20 | ···················· | ···················· | ···················· | ···················· | ······C············· | ···················· | |
| H | 6015 | hCoV-19/England/OXON-B4EA7/2020\|EPI_ISL_535020\|202 | ···················· | ···················· | ···················· | ···················· | ···················· | ···················· | |
| H | 6016 | hCoV-19/England/OXON-B4EC5/2020\|EPI_ISL_535021\|202 | ···················· | ···················· | ···················· | ···················· | ···················· | ···················· | |
| H | 6017 | hCoV-19/England/OXON-B4F4A/2020\|EPI_ISL_535022\|202 | ···················· | ···················· | ···················· | ···················· | ···················· | ···················· | |
| H | 6018 | hCoV-19/England/OXON-B4FB3/2020\|EPI_ISL_535023\|202 | ···················· | ···················· | ···················· | ···················· | ···················· | ···················· | |
| H | 6019 | hCoV-19/England/PORT-2D2005/2020\|EPI_ISL_535024\|20 | ···················· | ···················· | ···················· | ···················· | ···················· | ···················· | |
| H | 6020 | hCoV-19/England/PORT-2D208D/2020\|EPI_ISL_535025\|20 | ···················· | ···················· | ···················· | ···················· | ···················· | ···················· | |
| H | 6021 | hCoV-19/England/PORT-2D3417/2020\|EPI_ISL_535027\|20 | ···················· | ···················· | ···················· | ···················· | ···················· | ···················· | |
| H | 6022 | hCoV-19/England/PORT-2D3426/2020\|EPI_ISL_535028\|20 | ···················· | ···················· | ···················· | ···················· | ···················· | ···················· | |
| H | 6023 | hCoV-19/England/PORT-2D3444/2020\|EPI_ISL_535029\|20 | ···················· | ···················· | ···················· | ···················· | ···················· | ···················· | |
| H | 6024 | hCoV-19/England/PORT-2D3471/2020\|EPI_ISL_535031\|20 | ···················· | ···················· | ···················· | ···················· | ···················· | ···················· | |
| H | 6025 | hCoV-19/England/PORT-2D349F/2020\|EPI_ISL_535032\|20 | ···················· | ···················· | ···················· | ···················· | ···················· | ···················· | |
| H | 6026 | hCoV-19/England/PORT-2D3550/2020\|EPI_ISL_535036\|20 | ···················· | ···················· | ···················· | ···················· | ···················· | ···················· | |

| | | | | | | | | |
|---|---|---|---|---|---|---|---|---|
| H | 6027 hCoV-19/England/PORT-2D35D8/2020\|EPI_ISL_535038\|20 | .................... | .................... | .................... | .................... | .................... | .................... | |
| H | 6028 hCoV-19/England/SHEF-C0998/2020\|EPI_ISL_535043\|202 | .................... | .................... | .................... | .................... | .................... | .................... | |
| H | 6029 hCoV-19/England/SHEF-C0FE1/2020\|EPI_ISL_535044\|202 | .................... | .................... | .................... | .................... | .................... | .................... | |
| H | 6030 hCoV-19/England/SHEF-C1487/2020\|EPI_ISL_535047\|202 | .................... | .................... | .................... | .................... | .................... | .................... | |
| H | 6031 hCoV-19/England/SHEF-C3D8A/2020\|EPI_ISL_535051\|202 | .................... | .................... | .................... | .................... | .................... | .................... | |
| H | 6032 hCoV-19/England/SHEF-C4114/2020\|EPI_ISL_535052\|202 | .................... | .................... | .................... | .................... | .................... | .................... | |
| H | 6033 hCoV-19/England/SHEF-C51F5/2020\|EPI_ISL_535056\|202 | .................... | .................... | .................... | .................... | .................... | .................... | |
| H | 6034 hCoV-19/England/SHEF-C6419/2020\|EPI_ISL_535060\|202 | .................... | .................... | .................... | .................... | .................... | .................... | |
| H | 6035 hCoV-19/England/SHEF-C64DD/2020\|EPI_ISL_535061\|202 | .................... | .................... | .................... | .................... | .................... | .................... | |
| H | 6036 hCoV-19/England/SHEF-C6E93/2020\|EPI_ISL_535062\|202 | .................... | .................... | .................... | .................... | .................... | .................... | |
| H | 6037 hCoV-19/Canada/QC-LSPQ-L00243222/2020\|EPI_ISL_5363 | .................... | .................... | .................... | .................... | .................... | .................... | |
| H | 6038 hCoV-19/Canada/QC-LSPQ-L00243245/2020\|EPI_ISL_5363 | .................... | .................... | .................... | .................... | .................... | .................... | |
| H | 6039 hCoV-19/Canada/QC-LSPQ-L00243257/2020\|EPI_ISL_5363 | .................... | .................... | .................... | .................... | .................... | .................... | |
| H | 6040 hCoV-19/Canada/QC-LSPQ-L00243284/2020\|EPI_ISL_5363 | .................... | .................... | .................... | .................... | .................... | .................... | |
| H | 6041 hCoV-19/Canada/QC-LSPQ-L00243313/2020\|EPI_ISL_5363 | .................... | .................... | .................... | .................... | .................... | .................... | |
| H | 6042 hCoV-19/Canada/QC-LSPQ-L00243317/2020\|EPI_ISL_5363 | .................... | .................... | .................... | .................... | .................... | .................... | |
| H | 6043 hCoV-19/Canada/QC-LSPQ-L00243323/2020\|EPI_ISL_5363 | .................... | .................... | .................... | .................... | .................... | .................... | |
| H | 6044 hCoV-19/Canada/QC-LSPQ-L00243325/2020\|EPI_ISL_5363 | .................... | .................... | .................... | .................... | .................... | .................... | |
| H | 6045 hCoV-19/Canada/QC-LSPQ-L00243328/2020\|EPI_ISL_5363 | .................... | .................... | .................... | .................... | .................... | .................... | |
| H | 6046 hCoV-19/Canada/QC-LSPQ-L00243345/2020\|EPI_ISL_5363 | .................... | .................... | .................... | .................... | .................... | .................... | |
| H | 6047 hCoV-19/Canada/QC-LSPQ-L00243347/2020\|EPI_ISL_5363 | .................... | .................... | .................... | .................... | .................... | .................... | |
| H | 6048 hCoV-19/Canada/QC-LSPQ-L00243417/2020\|EPI_ISL_5363 | .................... | .................... | .................... | .................... | .................... | .................... | |
| H | 6049 hCoV-19/Canada/QC-LSPQ-L00243460/2020\|EPI_ISL_5363 | .................... | .................... | .................... | .................... | .................... | .................... | |
| H | 6050 hCoV-19/Canada/QC-LSPQ-L00243560/2020\|EPI_ISL_5363 | .................... | .................... | .................... | .................... | .................... | .................... | |
| H | 6051 hCoV-19/Canada/QC-LSPQ-L00243603/2020\|EPI_ISL_5363 | .................... | .................... | .................... | .................... | .................... | .................... | |
| H | 6052 hCoV-19/Canada/QC-LSPQ-L00243607/2020\|EPI_ISL_5363 | .................... | .................... | .................... | .................... | .................... | .................... | |
| H | 6053 hCoV-19/Canada/QC-LSPQ-L00243608/2020\|EPI_ISL_5363 | .................... | .................... | .................... | .................... | .................... | .................... | |
| H | 6054 hCoV-19/Canada/QC-LSPQ-L00243673/2020\|EPI_ISL_5363 | .................... | .................... | .................... | .................... | .................... | .................... | |
| H | 6055 hCoV-19/Canada/QC-LSPQ-L00243675/2020\|EPI_ISL_5363 | .................... | .................... | .................... | .................... | .................... | .................... | |
| H | 6056 hCoV-19/Canada/QC-LSPQ-L00243708/2020\|EPI_ISL_5363 | .................... | .................... | .................... | .................... | .................... | .................... | |
| H | 6057 hCoV-19/Canada/QC-LSPQ-L00243710/2020\|EPI_ISL_5363 | .................... | .................... | .................... | .................... | .................... | .................... | |
| H | 6058 hCoV-19/Canada/QC-LSPQ-L00243746/2020\|EPI_ISL_5363 | .................... | .................... | .................... | .................... | .................... | .................... | |
| H | 6059 hCoV-19/Canada/QC-LSPQ-L00243755/2020\|EPI_ISL_5363 | .................... | .................... | .................... | .................... | .................... | .................... | |
| H | 6060 hCoV-19/Canada/QC-LSPQ-L00243761/2020\|EPI_ISL_5363 | .................... | .................... | .................... | .................... | .................... | .................... | |
| H | 6061 hCoV-19/Canada/QC-LSPQ-L00243797/2020\|EPI_ISL_5363 | .................... | .................... | .................... | .................... | .................... | .................... | |
| H | 6062 hCoV-19/Canada/QC-LSPQ-L00243864/2020\|EPI_ISL_5363 | .................... | .................... | .................... | .................... | .................... | .................... | |
| H | 6063 hCoV-19/Canada/QC-LSPQ-L00243875/2020\|EPI_ISL_5363 | .................... | .................... | .................... | .................... | .................... | .................... | |
| H | 6064 hCoV-19/Canada/QC-LSPQ-L00243976/2020\|EPI_ISL_5363 | .................... | .................... | .................... | .................... | .................... | .................... | |
| H | 6065 hCoV-19/Canada/QC-LSPQ-L00243994/2020\|EPI_ISL_5363 | .................... | .................... | .................... | .................... | .................... | .................... | |
| H | 6066 hCoV-19/Canada/QC-LSPQ-L00244008/2020\|EPI_ISL_5363 | .................... | .................... | .................... | .................... | .................... | .................... | |
| H | 6067 hCoV-19/Canada/QC-LSPQ-L00244068/2020\|EPI_ISL_5363 | .................... | .................... | .................... | .................... | .................... | .................... | |
| H | 6068 hCoV-19/Canada/QC-LSPQ-L00244094/2020\|EPI_ISL_5363 | .................... | .................... | .................... | .................... | .................... | .................... | |
| H | 6069 hCoV-19/Canada/QC-LSPQ-L00244160/2020\|EPI_ISL_5363 | .................... | .................... | .................... | .................... | .................... | .................... | |
| H | 6070 hCoV-19/Canada/QC-LSPQ-L00244197/2020\|EPI_ISL_5363 | .................... | .................... | .................... | .................... | .................... | .................... | |
| H | 6071 hCoV-19/Canada/QC-LSPQ-L00244326/2020\|EPI_ISL_5363 | .................... | .................... | .................... | .................... | .................... | .................... | |
| H | 6072 hCoV-19/Canada/QC-LSPQ-L00244338/2020\|EPI_ISL_5363 | .................... | .................... | .................... | .................... | .................... | .................... | |
| H | 6073 hCoV-19/Canada/QC-LSPQ-L00244359/2020\|EPI_ISL_5363 | .................... | .................... | .................... | .................... | .................... | .................... | |
| H | 6074 hCoV-19/Spain/CN-IBV-97006476/2020\|EPI_ISL_537693\| | .................... | .................... | .................... | .................... | .................... | .................... | |
| H | 6076 hCoV-19/Italy/LOM-UniMI-L213/2020\|EPI_ISL_542200\|2 | .................... | .................... | .................... | .................... | .................... | .................... | |
| H | 6077 hCoV-19/Italy/LOM-UniMI-L214/2020\|EPI_ISL_542201\|2 | .................... | .................... | .................... | .................... | .................... | .................... | |
| H | 6078 hCoV-19/Italy/LOM-UniMI-L215/2020\|EPI_ISL_542202\|2 | .................... | .................... | .................... | .................... | .................... | .................... | |
| H | 6079 hCoV-19/Italy/LOM-UniMI-L216/2020\|EPI_ISL_542203\|2 | .................... | .................... | .................... | .................... | .................... | .................... | |
| H | 6080 hCoV-19/Italy/LOM-UniMI-L217/2020\|EPI_ISL_542204\|2 | .................... | .................... | .................... | .................... | .................... | .................... | |
| H | 6081 hCoV-19/Italy/LOM-UniMI-L218/2020\|EPI_ISL_542205\|2 | .................... | .................... | .................... | .................... | .................... | .................... | |
| H | 6082 hCoV-19/Italy/LOM-UniMI-L219/2020\|EPI_ISL_542206\|2 | .................... | .................... | .................... | .................... | .................... | .................... | |
| H | 6083 hCoV-19/Italy/LOM-UniMI-L220/2020\|EPI_ISL_542207\|2 | .................... | .................... | .................... | .................... | .................... | .................... | |
| H | 6084 hCoV-19/Italy/LOM-UniMI-L221/2020\|EPI_ISL_542208\|2 | .................... | .................... | .................... | .................... | .................... | .................... | |
| H | 6085 hCoV-19/Italy/LOM-UniMI-L222/2020\|EPI_ISL_542209\|2 | .................... | .................... | .................... | .................... | .................... | .................... | |
| H | 6086 hCoV-19/Italy/LOM-UniMI-L223/2020\|EPI_ISL_542210\|2 | .................... | .................... | .................... | .................... | .................... | .................... | |
| H | 6087 hCoV-19/Italy/LOM-UniMI-L224/2020\|EPI_ISL_542211\|2 | .................... | .................... | .................... | .................... | .................... | .................... | |
| H | 6088 hCoV-19/Italy/LOM-UniMI-L225/2020\|EPI_ISL_542212\|2 | .................... | .................... | .................... | .................... | .................... | .................... | |
| H | 6089 hCoV-19/Italy/LOM-UniMI-L226/2020\|EPI_ISL_542213\|2 | .................... | .................... | .................... | .................... | .................... | .................... | |
| H | 6090 hCoV-19/Italy/LOM-UniMI-L227/2020\|EPI_ISL_542214\|2 | .................... | .................... | .................... | .................... | .................... | .................... | |
| H | 6091 hCoV-19/Italy/LOM-UniMI-L228/2020\|EPI_ISL_542215\|2 | .................... | .................... | .................... | .................... | .................... | .................... | |
| H | 6092 hCoV-19/Italy/LOM-UniMI-L229/2020\|EPI_ISL_542216\|2 | .................... | .................... | .................... | .................... | .................... | .................... | |
| H | 6093 hCoV-19/Italy/LOM-UniMI-L230/2020\|EPI_ISL_542217\|2 | .................... | .................... | .................... | .................... | .................... | .................... | |
| H | 6094 hCoV-19/Italy/LOM-UniMI-L239/2020\|EPI_ISL_542218\|2 | .................... | .................... | .................... | .................... | .................... | .................... | |
| H | 6095 hCoV-19/Italy/LOM-UniMI-L240/2020\|EPI_ISL_542219\|2 | .................... | .................... | .................... | .................... | .................... | .................... | |
| H | 6096 hCoV-19/Italy/LOM-UniMI-L242/2020\|EPI_ISL_542220\|2 | .................... | .................... | .................... | .................... | .................... | .................... | |
| H | 6097 hCoV-19/Italy/LOM-UniMI-L249/2020\|EPI_ISL_542221\|2 | .................... | .................... | .................... | .................... | .................... | .................... | |
| H | 6098 hCoV-19/Italy/LOM-UniMI-L250/2020\|EPI_ISL_542222\|2 | .................... | .................... | .................... | .................... | .................... | .................... | |
| H | 6099 hCoV-19/Italy/LOM-UniMI-L251/2020\|EPI_ISL_542223\|2 | .................... | .................... | .................... | .................... | .................... | .................... | |
| H | 6100 hCoV-19/Italy/LOM-UniMI-L252/2020\|EPI_ISL_542224\|2 | .................... | .................... | .................... | .................... | .................... | .................... | |
| H | 6101 hCoV-19/USA/TX-HMH-4290/2020\|EPI_ISL_543500\|2020-0 | .................... | .................... | .................... | .................... | ...............N.... | NNN................. | 1 |
| H | 6102 hCoV-19/USA/TX-HMH-4292/2020\|EPI_ISL_543502\|2020-0 | .................... | .................... | .................... | .................... | .................... | .................... | |
| H | 6104 hCoV-19/USA/TX-HMH-4294/2020\|EPI_ISL_543504\|2020-0 | .................... | .................... | .................... | .................... | .................... | .................... | |
| H | 6105 hCoV-19/USA/TX-HMH-4296/2020\|EPI_ISL_543506\|2020-0 | .................... | .................... | .................... | .................... | .................... | .................... | |
| H | 6106 hCoV-19/USA/TX-HMH-4297/2020\|EPI_ISL_543507\|2020-0 | .................... | .................... | .................... | .................... | .................... | .................... | |
| H | 6107 hCoV-19/USA/TX-HMH-4299/2020\|EPI_ISL_543509\|2020-0 | .................... | .................... | .................... | .................... | .................... | .................... | |
| H | 6108 hCoV-19/USA/TX-HMH-4301/2020\|EPI_ISL_543511\|2020-0 | .................... | .................... | .................... | .................... | .................... | .................... | |

| | | | | | | | | |
|---|---|---|---|---|---|---|---|---|
| H | 6109 hCoV-19/USA/TX-HMH-4302/2020\|EPI_ISL_543512\|2020-0 | ···················· | ···················· | ···················· | ···················· | ···················· | ···················· | |
| H | 6110 hCoV-19/USA/TX-HMH-4303/2020\|EPI_ISL_543513\|2020-0 | ···················· | ···················· | ···················· | ···················· | ···················· | ···················· | |
| H | 6112 hCoV-19/USA/TX-HMH-4305/2020\|EPI_ISL_543515\|2020-0 | ···················· | ···················· | ···················· | ···················· | ···················· | ···················· | |
| H | 6113 hCoV-19/USA/TX-HMH-4309/2020\|EPI_ISL_543518\|2020-0 | ···················· | ···················· | ···················· | ···················· | ···················· | ···················· | |
| H | 6114 hCoV-19/USA/TX-HMH-4310/2020\|EPI_ISL_543519\|2020-0 | ···················· | ···················· | ···················· | ···················· | ···················· | ···················· | |
| H | 6116 hCoV-19/USA/TX-HMH-4312/2020\|EPI_ISL_543521\|2020-0 | ···················· | ···················· | ···················· | ···················· | ···················· | ···················· | |
| H | 6118 hCoV-19/USA/TX-HMH-4315/2020\|EPI_ISL_543524\|2020-0 | ···················· | ···················· | ···················· | ···················· | ···················· | ···················· | |
| H | 6119 hCoV-19/USA/TX-HMH-4316/2020\|EPI_ISL_543525\|2020-0 | ···················· | ·A·················· | ···················· | ···················· | ···················· | ···················· | |
| H | 6120 hCoV-19/USA/TX-HMH-4318/2020\|EPI_ISL_543527\|2020-0 | ···················· | ···················· | ···················· | ···················· | ···················· | ···················· | |
| H | 6122 hCoV-19/USA/TX-HMH-4320/2020\|EPI_ISL_543529\|2020-0 | ···················· | ···················· | ···················· | ···················· | ···················· | ···················· | |
| H | 6123 hCoV-19/USA/TX-HMH-4321/2020\|EPI_ISL_543530\|2020-0 | ···················· | ···················· | ···················· | ···················· | ···················· | ···················· | |
| H | 6125 hCoV-19/USA/TX-HMH-4324/2020\|EPI_ISL_543533\|2020-0 | ···················· | NN·····N············ | ···················· | ···················· | ···················· | ···················· | 1 |
| H | 6126 hCoV-19/USA/TX-HMH-4325/2020\|EPI_ISL_543534\|2020-0 | ···················· | ···················· | ···················· | ···················· | ···················· | ···················· | |
| H | 6129 hCoV-19/USA/TX-HMH-4328/2020\|EPI_ISL_543537\|2020-0 | ···················· | ···················· | ···················· | ···················· | ···················· | ···················· | |
| H | 6130 hCoV-19/USA/TX-HMH-4330/2020\|EPI_ISL_543538\|2020-0 | ···················· | ···················· | ···················· | ···················· | ···················· | ···················· | |
| H | 6131 hCoV-19/USA/TX-HMH-4332/2020\|EPI_ISL_543540\|2020-0 | ···················· | ···················· | ···················· | ···················· | ···················· | ···················· | |
| H | 6132 hCoV-19/USA/TX-HMH-4334/2020\|EPI_ISL_543541\|2020-0 | ···················· | ···················· | ···················· | ···················· | ···················· | ···················· | |
| H | 6134 hCoV-19/USA/TX-HMH-4341/2020\|EPI_ISL_543544\|2020-0 | ···················· | ···················· | ···················· | ···················· | ···················· | ···················· | |
| H | 6135 hCoV-19/USA/TX-HMH-4342/2020\|EPI_ISL_543545\|2020-0 | ···················· | ···················· | ···················· | ···················· | ···················· | ··A················· | |
| H | 6136 hCoV-19/USA/TX-HMH-4343/2020\|EPI_ISL_543546\|2020-0 | ···················· | ·············A······ | ···················· | ···················· | ···················· | ···················· | |
| H | 6137 hCoV-19/USA/TX-HMH-4346/2020\|EPI_ISL_543547\|2020-0 | ···················· | ···················· | ···················· | ···················· | ···················· | ···················· | |
| H | 6138 hCoV-19/USA/TX-HMH-4348/2020\|EPI_ISL_543549\|2020-0 | ···················· | ···················· | ···················· | ···················· | ···················· | ···················· | |
| H | 6139 hCoV-19/USA/TX-HMH-4349/2020\|EPI_ISL_543550\|2020-0 | ···················· | ·A·················· | ···················· | ···················· | ···················· | ······NN············ | |
| H | 6140 hCoV-19/USA/TX-HMH-4350/2020\|EPI_ISL_543551\|2020-0 | ···················· | ···················· | ···················· | ···················· | ···················· | ···················· | |
| H | 6141 hCoV-19/USA/TX-HMH-4351/2020\|EPI_ISL_543552\|2020-0 | ···················· | ···················· | ···················· | ···················· | ···················· | ···················· | |
| H | 6142 hCoV-19/USA/TX-HMH-4352/2020\|EPI_ISL_543553\|2020-0 | ···················· | ···················· | ···················· | ···················· | ···················· | ···················· | |
| H | 6144 hCoV-19/USA/TX-HMH-4356/2020\|EPI_ISL_543555\|2020-0 | ···················· | ···················· | ···················· | ···················· | ···················· | ···················· | |
| H | 6145 hCoV-19/England/QEUH-96B43B/2020\|EPI_ISL_531338\|20 | ···················· | ···················· | ···················· | ···················· | ···················· | ···················· | |
| H | 6146 hCoV-19/England/QEUH-96BDA9/2020\|EPI_ISL_531339\|20 | ···················· | ···················· | ···················· | ···················· | ···················· | ···················· | |
| H | 6147 hCoV-19/Northern Ireland/QEUH-96AB28/2020\|EPI_ISL_ | ···················· | ···················· | ···················· | ···················· | ···················· | ···················· | |
| H | 6148 hCoV-19/England/QEUH-96BE1F/2020\|EPI_ISL_531341\|20 | ···················· | ···················· | ···················· | ···················· | ···················· | ···················· | |
| H | 6149 hCoV-19/Northern Ireland/QEUH-96AC07/2020\|EPI_ISL_ | ···················· | ···················· | ···················· | ···················· | ···················· | ···················· | |
| H | 6150 hCoV-19/England/QEUH-96AC16/2020\|EPI_ISL_531343\|20 | ···················· | ···················· | ···················· | ···················· | ···················· | ···················· | |
| H | 6151 hCoV-19/England/QEUH-96B705/2020\|EPI_ISL_531344\|20 | ···················· | ···················· | ···················· | ···················· | ···················· | ···················· | |
| H | 6152 hCoV-19/England/QEUH-96BB90/2020\|EPI_ISL_531345\|20 | ···················· | ···················· | ···················· | ···················· | ···················· | ···················· | |
| H | 6153 hCoV-19/England/QEUH-96BC15/2020\|EPI_ISL_531346\|20 | ···················· | ···················· | ···················· | ···················· | ···················· | ···················· | |
| H | 6154 hCoV-19/England/QEUH-96AD04/2020\|EPI_ISL_531347\|20 | ···················· | ···················· | ···················· | ···················· | ···················· | ···················· | |
| H | 6155 hCoV-19/England/QEUH-96B037/2020\|EPI_ISL_531348\|20 | ···················· | ···················· | ···················· | ···················· | ···················· | ···················· | |
| H | 6156 hCoV-19/England/QEUH-96B19E/2020\|EPI_ISL_531349\|20 | ···················· | ···················· | ···················· | ···················· | ···················· | ···················· | |
| H | 6157 hCoV-19/England/QEUH-96B723/2020\|EPI_ISL_531350\|20 | ···················· | ···················· | ···················· | ···················· | ···················· | ···················· | |
| H | 6158 hCoV-19/England/QEUH-96B8A8/2020\|EPI_ISL_531351\|20 | ···················· | ···················· | ···················· | ···················· | ···················· | ···················· | |
| H | 6159 hCoV-19/England/QEUH-96BE6A/2020\|EPI_ISL_531353\|20 | ···················· | ···················· | ···················· | ···················· | ···················· | ···················· | |
| H | 6160 hCoV-19/England/QEUH-96B34D/2020\|EPI_ISL_531354\|20 | ···················· | ···················· | ···················· | ···················· | ···················· | ···················· | |
| H | 6161 hCoV-19/England/QEUH-96B477/2020\|EPI_ISL_531356\|20 | ···················· | ···················· | ···················· | ···················· | ···················· | ···················· | |
| H | 6162 hCoV-19/England/QEUH-96BBAF/2020\|EPI_ISL_531358\|20 | ···················· | ···················· | ···················· | ···················· | ···················· | ···················· | |
| H | 6163 hCoV-19/England/QEUH-8AACC0/2020\|EPI_ISL_532666\|20 | ···················· | ···················· | ···················· | ···················· | ···················· | ···················· | |
| H | 6164 hCoV-19/England/QEUH-8ACBB2/2020\|EPI_ISL_532667\|20 | ···················· | ···················· | ···················· | ···················· | ·········T·········· | ···················· | |
| H | 6165 hCoV-19/England/QEUH-8A6514/2020\|EPI_ISL_532668\|20 | ···················· | ···················· | ···················· | ···················· | ···················· | ···················· | |
| H | 6166 hCoV-19/Scotland/QEUH-89B864/2020\|EPI_ISL_532670\|2 | ···················· | ···················· | ···················· | ···················· | ···················· | ···················· | |
| H | 6167 hCoV-19/England/QEUH-894D5D/2020\|EPI_ISL_532671\|20 | ···················· | ···················· | ···················· | ···················· | ···················· | ···················· | |
| H | 6168 hCoV-19/Northern Ireland/QEUH-8802A0/2020\|EPI_ISL_ | ···················· | ···················· | ···················· | ···················· | ···················· | ···················· | |
| H | 6169 hCoV-19/England/QEUH-881BC2/2020\|EPI_ISL_532675\|20 | ···················· | ···················· | ···················· | ···················· | ···················· | ···················· | |
| H | 6170 hCoV-19/Northern Ireland/QEUH-8802BF/2020\|EPI_ISL_ | ···················· | ···················· | ···················· | ···················· | ···················· | ···················· | |
| H | 6171 hCoV-19/England/QEUH-8A6BD6/2020\|EPI_ISL_532677\|20 | ···················· | ···················· | ···················· | ···················· | ···················· | ···················· | |
| H | 6172 hCoV-19/England/QEUH-87F611/2020\|EPI_ISL_532678\|20 | ···················· | ···················· | ···················· | ···················· | ···················· | ···················· | |
| H | 6173 hCoV-19/England/QEUH-8AFCE9/2020\|EPI_ISL_532679\|20 | ···················· | ···················· | ···················· | ···················· | ···················· | ···················· | |
| H | 6174 hCoV-19/England/QEUH-885663/2020\|EPI_ISL_532680\|20 | ···················· | ···················· | ···················· | ···················· | ···················· | ···················· | |
| H | 6175 hCoV-19/England/QEUH-87DB41/2020\|EPI_ISL_532681\|20 | ···················· | ···················· | ···················· | ···················· | ···················· | ···················· | |
| H | 6176 hCoV-19/England/QEUH-89873D/2020\|EPI_ISL_532682\|20 | ···················· | ···················· | ···················· | ···················· | ···················· | ···················· | |
| H | 6177 hCoV-19/Scotland/QEUH-89AAD8/2020\|EPI_ISL_532683\|2 | ···················· | ···················· | ···················· | ···················· | ···················· | ···················· | |
| H | 6178 hCoV-19/England/QEUH-88A80E/2020\|EPI_ISL_532685\|20 | ···················· | ···················· | ···················· | ···················· | ···················· | ···················· | |
| H | 6179 hCoV-19/England/QEUH-882A88/2020\|EPI_ISL_532686\|20 | ···················· | ···················· | ···················· | ···················· | ···················· | ···················· | |
| H | 6180 hCoV-19/Northern Ireland/QEUH-87CC03/2020\|EPI_ISL_ | ···················· | ···················· | ···················· | ···················· | ···················· | ···················· | |
| H | 6181 hCoV-19/Scotland/QEUH-89BBD4/2020\|EPI_ISL_532689\|2 | ···················· | ···················· | ···················· | ···················· | ···················· | ···················· | |
| H | 6182 hCoV-19/England/QEUH-894CE7/2020\|EPI_ISL_532800\|20 | ···················· | ···················· | ···················· | ···················· | ···················· | ···················· | |
| H | 6183 hCoV-19/England/QEUH-896F37/2020\|EPI_ISL_532802\|20 | ···················· | ···················· | ···················· | ···················· | ···················· | ···················· | |
| H | 6184 hCoV-19/England/QEUH-8930A0/2020\|EPI_ISL_532803\|20 | ···················· | ···················· | ···················· | ···················· | ···················· | ···················· | |
| H | 6185 hCoV-19/Northern Ireland/QEUH-88932C/2020\|EPI_ISL_ | ···················· | ···················· | ···················· | ···················· | ···················· | ···················· | |
| H | 6186 hCoV-19/England/QEUH-896210/2020\|EPI_ISL_532805\|20 | ···················· | ···················· | ···················· | ···················· | ···················· | ···················· | |
| H | 6187 hCoV-19/England/QEUH-8AC305/2020\|EPI_ISL_532806\|20 | ···················· | ···················· | ···················· | ···················· | ···················· | ···················· | |
| H | 6188 hCoV-19/England/QEUH-881403/2020\|EPI_ISL_532807\|20 | ···················· | ·A·················· | ···················· | ···················· | ···················· | ···················· | |
| H | 6189 hCoV-19/England/QEUH-87FDB2/2020\|EPI_ISL_532808\|20 | ···················· | ···················· | ···················· | ···················· | ···················· | ···················· | |
| H | 6190 hCoV-19/England/QEUH-884D8D/2020\|EPI_ISL_532809\|20 | ···················· | ···················· | ···················· | ···················· | ···················· | ···················· | |
| H | 6191 hCoV-19/England/QEUH-89D729/2020\|EPI_ISL_532810\|20 | ···················· | ···················· | ···················· | ···················· | ···················· | ···················· | |
| H | 6192 hCoV-19/Scotland/QEUH-89C0D4/2020\|EPI_ISL_532811\|2 | ···················· | ···················· | ···················· | ···················· | ···················· | ···················· | |
| H | 6193 hCoV-19/England/QEUH-897622/2020\|EPI_ISL_532812\|20 | ···················· | ···················· | ···················· | ···················· | ···················· | ···················· | |
| H | 6194 hCoV-19/England/QEUH-899383/2020\|EPI_ISL_532813\|20 | ···················· | ···················· | ···················· | ···················· | ···················· | ···················· | |
| H | 6195 hCoV-19/England/QEUH-892E89/2020\|EPI_ISL_532815\|20 | ···················· | ···················· | ···················· | ···················· | ···················· | ···················· | |
| H | 6196 hCoV-19/England/QEUH-89EBA4/2020\|EPI_ISL_532816\|20 | ···················· | ···················· | ···················· | ···················· | ···················· | ···················· | |
| H | 6197 hCoV-19/England/QEUH-88CEDD/2020\|EPI_ISL_532817\|20 | ···················· | ···················· | ···················· | ···················· | ···················· | ···················· | |

| | | | | | | | |
|---|---|---|---|---|---|---|---|
| H | 6198 hCoV-19/Wales/QEUH-8ADF3D/2020\|EPI_ISL_532819\|2020 | ···················· | ···················· | ···················· | ···················· | ···················· | ···················· |
| H | 6199 hCoV-19/England/SHEF-C73EE/2020\|EPI_ISL_535063\|202 | ···················· | ···················· | ···················· | ···················· | ···················· | ···················· |
| H | 6200 hCoV-19/England/SHEF-C8B7C/2020\|EPI_ISL_535064\|202 | ···················· | ···················· | ···················· | ···················· | ···················· | ···················· |
| H | 6201 hCoV-19/England/SHEF-C98B0/2020\|EPI_ISL_535066\|202 | ···················· | ···················· | ···················· | ···················· | ···················· | ···················· |
| H | 6202 hCoV-19/England/SHEF-C9FAC/2020\|EPI_ISL_535069\|202 | ···················· | ···················· | ···T················ | ···················· | ···················· | ···················· |
| H | 6203 hCoV-19/England/SHEF-CAC3B/2020\|EPI_ISL_535071\|202 | ···················· | ···················· | ···················· | ···················· | ···················· | ···················· |
| H | 6204 hCoV-19/England/SHEF-CBAD6/2020\|EPI_ISL_535073\|202 | ···················· | ···················· | ···················· | ···················· | ···················· | ···················· |
| H | 6205 hCoV-19/England/SHEF-CC273/2020\|EPI_ISL_535075\|202 | ···················· | ···················· | ···················· | ···················· | ···················· | ···················· |
| H | 6206 hCoV-19/England/SHEF-CC96F/2020\|EPI_ISL_535077\|202 | ···················· | ···················· | ···················· | ···················· | ···················· | ···················· |
| H | 6207 hCoV-19/England/SHEF-CCDEB/2020\|EPI_ISL_535079\|202 | ···················· | ···················· | ···················· | ···················· | ···················· | ···················· |
| H | 6208 hCoV-19/England/SHEF-CD9D7/2020\|EPI_ISL_535080\|202 | ···················· | ···················· | ···················· | ···················· | ···················· | ···················· |
| H | 6209 hCoV-19/England/SHEF-CDD62/2020\|EPI_ISL_535081\|202 | ···················· | ···················· | ···················· | ···················· | ···················· | ···················· |
| H | 6210 hCoV-19/England/SHEF-CDE14/2020\|EPI_ISL_535082\|202 | ···················· | ···················· | ···················· | ···················· | ···················· | ···················· |
| H | 6211 hCoV-19/England/SHEF-CE11A/2020\|EPI_ISL_535084\|202 | ···················· | ···················· | ···················· | ···················· | ···················· | ···················· |
| H | 6212 hCoV-19/England/SHEF-CE39C/2020\|EPI_ISL_535085\|202 | ···················· | ···················· | ···················· | ···················· | ···················· | ···················· |
| H | 6213 hCoV-19/England/SHEF-CEBEF/2020\|EPI_ISL_535087\|202 | ···················· | ···················· | ···················· | ···················· | ···················· | ···················· |
| H | 6214 hCoV-19/England/SHEF-CF137/2020\|EPI_ISL_535088\|202 | ···················· | ···················· | ···················· | ···················· | ···················· | ···················· |
| H | 6215 hCoV-19/England/SHEF-CF79F/2020\|EPI_ISL_535089\|202 | ···················· | ···················· | ···················· | ···················· | ···················· | ···················· |
| H | 6216 hCoV-19/England/SHEF-D02C7/2020\|EPI_ISL_535091\|202 | ···················· | ···················· | ···················· | ···················· | ···················· | ···················· |
| H | 6217 hCoV-19/England/SHEF-D1B25/2020\|EPI_ISL_535095\|202 | ···················· | ···················· | ···················· | ···················· | ···················· | ···················· |
| H | 6218 hCoV-19/England/SHEF-D1CC8/2020\|EPI_ISL_535096\|202 | ···················· | ···················· | ···················· | ···················· | ···················· | ···················· |
| H | 6219 hCoV-19/England/SHEF-D1EB3/2020\|EPI_ISL_535097\|202 | ···················· | ···················· | ···················· | ···················· | ···················· | ···················· |
| H | 6220 hCoV-19/England/SHEF-D2456/2020\|EPI_ISL_535099\|202 | ···················· | ···················· | ···················· | ···················· | ···················· | ···················· |
| H | 6221 hCoV-19/Canada/QC-LSPQ-L00237930/2020\|EPI_ISL_5362 | ···················· | ···················· | ···················· | ···················· | ···················· | ···················· |
| H | 6222 hCoV-19/Canada/QC-LSPQ-L00237942/2020\|EPI_ISL_5362 | ···················· | ···················· | ···················· | ···················· | ···················· | ···················· |
| H | 6223 hCoV-19/Canada/QC-LSPQ-L00237972/2020\|EPI_ISL_5362 | ···················· | ···················· | ···················· | ···················· | ···················· | ···················· |
| H | 6224 hCoV-19/Canada/QC-LSPQ-L00238226/2020\|EPI_ISL_5362 | ···················· | ···················· | ···················· | ···················· | ···················· | ···················· |
| H | 6225 hCoV-19/Canada/QC-LSPQ-L00238246/2020\|EPI_ISL_5362 | ···················· | ···················· | ···················· | ···················· | ···················· | ···················· |
| H | 6226 hCoV-19/Canada/QC-LSPQ-L00238342/2020\|EPI_ISL_5362 | ···················· | ···················· | ···················· | ···················· | ···················· | ···················· |
| H | 6227 hCoV-19/Canada/QC-LSPQ-L00238445/2020\|EPI_ISL_5362 | ···················· | ···················· | ···················· | ···················· | ···················· | ···················· |
| H | 6228 hCoV-19/Canada/QC-LSPQ-L00238518/2020\|EPI_ISL_5362 | ···················· | ···················· | ···················· | ···················· | ···················· | ···················· |
| H | 6229 hCoV-19/Canada/QC-LSPQ-L00238674/2020\|EPI_ISL_5362 | ···················· | ···················· | ···················· | ···················· | ···················· | ···················· |
| H | 6230 hCoV-19/Canada/QC-LSPQ-L00238675/2020\|EPI_ISL_5362 | ···················· | ···················· | ···················· | ···················· | ···················· | ···················· |
| H | 6231 hCoV-19/Canada/QC-LSPQ-L00238722/2020\|EPI_ISL_5362 | ···················· | ···················· | ···················· | ···················· | ···················· | ···················· |
| H | 6232 hCoV-19/Canada/QC-LSPQ-L00238725/2020\|EPI_ISL_5362 | ···················· | ···················· | ···················· | ···················· | ···················· | ···················· |
| H | 6233 hCoV-19/Canada/QC-LSPQ-L00238793/2020\|EPI_ISL_5362 | ···················· | ···················· | ···················· | ···················· | ···················· | ···················· |
| H | 6234 hCoV-19/Canada/QC-LSPQ-L00238797/2020\|EPI_ISL_5362 | ···················· | ···················· | ···················· | ···················· | ···················· | ···················· |
| H | 6235 hCoV-19/Canada/QC-LSPQ-L00238803/2020\|EPI_ISL_5362 | ···················· | ···················· | ···················· | ···················· | ···················· | ···················· |
| H | 6236 hCoV-19/Canada/QC-LSPQ-L00238819/2020\|EPI_ISL_5362 | ···················· | ···················· | ···················· | ···················· | ···················· | ···················· |
| H | 6237 hCoV-19/Canada/QC-LSPQ-L00238862/2020\|EPI_ISL_5362 | ···················· | ···················· | ···················· | ···················· | ···················· | ···················· |
| H | 6238 hCoV-19/Canada/QC-LSPQ-L00238888/2020\|EPI_ISL_5362 | ···················· | ···················· | ···················· | ···················· | ···················· | ···················· |
| H | 6239 hCoV-19/Canada/QC-LSPQ-L00238922/2020\|EPI_ISL_5362 | ···················· | ···················· | ···················· | ···················· | ···················· | ···················· |
| H | 6240 hCoV-19/Canada/QC-LSPQ-L00238978/2020\|EPI_ISL_5362 | ···················· | ···················· | ···················· | ···················· | ···················· | ···················· |
| H | 6241 hCoV-19/Canada/QC-LSPQ-L00244354/2020\|EPI_ISL_5363 | ···················· | ···················· | ···················· | ···················· | ···················· | ···················· |
| H | 6242 hCoV-19/Lithuania/MR-LUHS-Eilnr97/2020\|EPI_ISL_536 | ···················· | ···················· | ···················· | ···················· | ···················· | ···················· |
| H | 6243 hCoV-19/Colombia/ANT-UdeA-200325-01/2020\|EPI_ISL_5 | ···················· | ···················· | ···················· | ···················· | ···················· | ···················· |
| H | 6244 hCoV-19/USA/CA-UCLA-160/2020\|EPI_ISL_537500\|2020-0 | ···················· | ···················· | ···················· | ···················· | ···················· | ···················· |
| H | 6245 hCoV-19/USA/CA-UCLA-162/2020\|EPI_ISL_537501\|2020-0 | ···················· | ···················· | ···················· | ···················· | ···················· | ···················· |
| H | 6246 hCoV-19/USA/CA-UCLA-163/2020\|EPI_ISL_537502\|2020-0 | ···················· | ···················· | ···················· | ···················· | ···················· | ···················· |
| H | 6247 hCoV-19/USA/CA-UCLA-164/2020\|EPI_ISL_537503\|2020-0 | ···················· | ···················· | ···················· | ···················· | ···················· | ···················· |
| H | 6248 hCoV-19/USA/CA-UCLA-165/2020\|EPI_ISL_537504\|2020-0 | ···················· | ···················· | ···················· | ···················· | ···················· | ···················· |
| H | 6249 hCoV-19/USA/CA-UCLA-17/2020\|EPI_ISL_537505\|2020-04 | ···················· | ···················· | ···················· | ···················· | ···················· | ···················· |
| H | 6250 hCoV-19/USA/CA-UCLA-18/2020\|EPI_ISL_537506\|2020-05 | ···T················ | ···················· | ···················· | ···················· | ···················· | ···················· |
| H | 6251 hCoV-19/USA/CA-UCLA-180/2020\|EPI_ISL_537507\|2020-0 | ···················· | ···················· | ···················· | ···················· | ···················· | ···················· |
| H | 6252 hCoV-19/USA/CA-UCLA-19/2020\|EPI_ISL_537508\|2020-05 | ···················· | ···················· | ···················· | ···················· | ···················· | ···················· |
| H | 6253 hCoV-19/USA/CA-UCLA-2/2020\|EPI_ISL_537509\|2020-04- | ···················· | ···················· | ···················· | ···················· | ···················· | ···················· |
| H | 6254 hCoV-19/USA/CA-UCLA-20/2020\|EPI_ISL_537510\|2020-05 | ···················· | ···················· | ···················· | ···················· | ···················· | ···················· |
| H | 6255 hCoV-19/USA/CA-UCLA-202/2020\|EPI_ISL_537512\|2020-0 | ···················· | ···················· | ···················· | ···················· | ···················· | ···················· |
| H | 6256 hCoV-19/USA/CA-UCLA-206/2020\|EPI_ISL_537513\|2020-0 | ···················· | ···················· | ···················· | ···················· | ···················· | ···················· |
| H | 6257 hCoV-19/USA/CA-UCLA-207/2020\|EPI_ISL_537514\|2020-0 | ···················· | ···················· | ···················· | ···················· | ···················· | ···················· |
| H | 6258 hCoV-19/USA/CA-UCLA-208/2020\|EPI_ISL_537515\|2020-0 | ···················· | ···················· | ···················· | ···················· | ···················· | ···················· |
| H | 6259 hCoV-19/USA/CA-UCLA-21/2020\|EPI_ISL_537517\|2020-05 | ···················· | ···················· | ···················· | ···················· | ···················· | ···················· |
| H | 6260 hCoV-19/USA/CA-UCLA-216/2020\|EPI_ISL_537523\|2020-0 | ···················· | ···················· | ···················· | ···················· | ···················· | ···················· |
| H | 6261 hCoV-19/USA/CA-UCLA-219/2020\|EPI_ISL_537525\|2020-0 | ···················· | ···················· | ···················· | ···················· | ···················· | ···················· |
| H | 6262 hCoV-19/USA/CA-UCLA-22/2020\|EPI_ISL_537526\|2020-05 | ···················· | ···················· | ···················· | ···················· | ···················· | ···················· |
| H | 6263 hCoV-19/USA/CA-UCLA-220/2020\|EPI_ISL_537527\|2020-0 | ···················· | ···················· | ···················· | ···················· | ···················· | ···················· |
| H | 6264 hCoV-19/USA/CA-UCLA-221/2020\|EPI_ISL_537528\|2020-0 | ···················· | ···················· | ···················· | ···················· | ···················· | ···················· |
| H | 6265 hCoV-19/USA/CA-UCLA-222/2020\|EPI_ISL_537529\|2020-0 | ···················· | ·A·················· | ···················· | ···················· | ···················· | ···················· |
| H | 6266 hCoV-19/USA/CA-UCLA-223/2020\|EPI_ISL_537530\|2020-0 | ···················· | ···················· | ···················· | ···················· | ···················· | ···················· |
| H | 6267 hCoV-19/USA/CA-UCLA-224/2020\|EPI_ISL_537531\|2020-0 | ···················· | ···················· | ···················· | ···················· | ···················· | ···················· |
| H | 6268 hCoV-19/USA/CA-UCLA-227/2020\|EPI_ISL_537532\|2020-0 | ···················· | ···················· | ···················· | ···················· | ···················· | ···················· |
| H | 6269 hCoV-19/USA/CA-UCLA-228/2020\|EPI_ISL_537533\|2020-0 | ···················· | ···················· | ···················· | ···················· | ···················· | ···················· |
| H | 6270 hCoV-19/USA/CA-UCLA-23/2020\|EPI_ISL_537535\|2020-05 | ···················· | ···················· | ···················· | ···················· | ···················· | ···················· |
| H | 6271 hCoV-19/USA/CA-UCLA-231/2020\|EPI_ISL_537537\|2020-0 | ···················· | ···················· | ···················· | ···················· | ···················· | ···················· |
| H | 6272 hCoV-19/USA/CA-UCLA-232/2020\|EPI_ISL_537538\|2020-0 | ···················· | ···················· | ···················· | ···················· | ···················· | ···················· |
| H | 6273 hCoV-19/USA/CA-UCLA-235/2020\|EPI_ISL_537541\|2020-0 | ···················· | ·A·················· | ···················· | ···················· | ···················· | ···················· |
| H | 6274 hCoV-19/USA/CA-UCLA-236/2020\|EPI_ISL_537542\|2020-0 | ···················· | ···················· | ···················· | ···················· | ···················· | ···················· |
| H | 6275 hCoV-19/USA/CA-UCLA-24/2020\|EPI_ISL_537545\|2020-05 | ···················· | ···················· | ···················· | ···················· | ···················· | ···················· |
| H | 6276 hCoV-19/USA/CA-UCLA-240/2020\|EPI_ISL_537546\|2020-0 | ···················· | ···················· | ···················· | ···················· | ···················· | ···················· |
| H | 6277 hCoV-19/USA/CA-UCLA-241/2020\|EPI_ISL_537547\|2020-0 | ···················· | ···················· | ···················· | ···················· | ···················· | ···················· |

| | | | | | | | | | |
|---|---|---|---|---|---|---|---|---|---|
| H | 6278 | hCoV-19/USA/CA-UCLA-25/2020\|EPI_ISL_537551\|2020-05 | ···················· | ···················· | ····················· | ···················· | ···················· | ···················· | |
| H | 6279 | hCoV-19/USA/CA-UCLA-252/2020\|EPI_ISL_537554\|2020-0 | ···················· | ·A·················· | ····················· | ···················· | ···················· | ···················· | |
| H | 6280 | hCoV-19/USA/CA-UCLA-26/2020\|EPI_ISL_537557\|2020-05 | ···················· | ···················· | ····················· | ···················· | ···················· | ···················· | |
| H | 6281 | hCoV-19/USA/CA-UCLA-260/2020\|EPI_ISL_537558\|2020-0 | ···················· | ···················· | ····················· | ···················· | ···················· | ···················· | |
| H | 6282 | hCoV-19/USA/CA-UCLA-27/2020\|EPI_ISL_537559\|2020-05 | ···················· | ···················· | ····················· | ···················· | ···················· | ···················· | |
| H | 6283 | hCoV-19/England/LEED-2AA3A3/2020\|EPI_ISL_538804\|20 | ···················· | ···················· | ····················· | ···················· | ···················· | ···················· | |
| H | 6284 | hCoV-19/England/LEED-2AA622/2020\|EPI_ISL_538805\|20 | ···················· | ···················· | ····················· | ···················· | ···················· | ···················· | |
| H | 6285 | hCoV-19/England/LEED-2A9A90/2020\|EPI_ISL_538806\|20 | ···················· | ···················· | ····················· | ···················· | ···················· | ···················· | |
| H | 6286 | hCoV-19/England/LEED-2AA1A9/2020\|EPI_ISL_538807\|20 | ···················· | ·A·················· | ····················· | ···················· | ···················· | ···················· | |
| H | 6287 | hCoV-19/England/LEED-2A9A27/2020\|EPI_ISL_538808\|20 | ···················· | ···················· | ····················· | ···················· | ···················· | ···················· | |
| H | 6288 | hCoV-19/England/LEED-2A9AAF/2020\|EPI_ISL_538809\|20 | ···················· | ··············C····· | ···········Y········· | ···················· | ···················· | ···················· | |
| H | 6289 | hCoV-19/England/LEED-2A94DE/2020\|EPI_ISL_538810\|20 | ···················· | ···················· | ····················· | ···················· | ···················· | ···················· | |
| H | 6290 | hCoV-19/England/LEED-2A93E0/2020\|EPI_ISL_538812\|20 | ···················· | ···A················ | ····················· | ···················· | ···················· | ···················· | |
| H | 6291 | hCoV-19/England/LEED-2A9BD9/2020\|EPI_ISL_538813\|20 | ···················· | ···················· | ····················· | ···················· | ···················· | ···················· | |
| H | 6292 | hCoV-19/England/LEED-2AA561/2020\|EPI_ISL_538814\|20 | ···················· | ···················· | ····················· | ···················· | ···················· | ···················· | |
| H | 6293 | hCoV-19/England/LEED-2AA1C7/2020\|EPI_ISL_538815\|20 | ···················· | ···················· | ····················· | ···················· | ···················· | ···················· | |
| H | 6294 | hCoV-19/England/LEED-2AA394/2020\|EPI_ISL_538816\|20 | ···················· | ···················· | ····················· | ···················· | ···················· | ···················· | |
| H | 6295 | hCoV-19/England/LEED-2A95AE/2020\|EPI_ISL_538817\|20 | ···················· | ···················· | ····················· | ···················· | ···················· | ···················· | |
| H | 6296 | hCoV-19/England/LEED-2AA367/2020\|EPI_ISL_538819\|20 | ···················· | ···················· | ····················· | ···················· | ···················· | ···················· | |
| H | 6297 | hCoV-19/England/LEED-2AA5AD/2020\|EPI_ISL_538820\|20 | ···················· | ···················· | ····················· | ···················· | ···················· | ···················· | |
| H | 6298 | hCoV-19/England/LEED-2AA604/2020\|EPI_ISL_538821\|20 | ···················· | ···················· | ····················· | ···················· | ···················· | ···················· | |
| H | 6299 | hCoV-19/England/LEED-2AA0F7/2020\|EPI_ISL_538822\|20 | ···················· | ···················· | ····················· | ···················· | ···················· | ···················· | |
| H | 6300 | hCoV-19/England/LEED-2AA15E/2020\|EPI_ISL_538823\|20 | ···················· | ···················· | ····················· | ···················· | ···················· | ···················· | |
| H | 6301 | hCoV-19/England/LEED-2AA21F/2020\|EPI_ISL_538824\|20 | ···················· | ···················· | ····················· | ···················· | ···················· | ···················· | |
| H | 6302 | hCoV-19/England/LEED-2AA428/2020\|EPI_ISL_538825\|20 | ···················· | ·A·················· | ····················· | ···················· | ···················· | ···················· | |
| H | 6303 | hCoV-19/England/LEED-2AA552/2020\|EPI_ISL_538826\|20 | ···················· | ·A·················· | ····················· | ···················· | ···················· | ···················· | |
| H | 6304 | hCoV-19/England/LEED-2AA26A/2020\|EPI_ISL_538827\|20 | ···················· | ···················· | ····················· | ···················· | ···················· | ···················· | |
| H | 6305 | hCoV-19/England/LEED-2A99EE/2020\|EPI_ISL_538828\|20 | ···················· | ···················· | ····················· | ···················· | ···················· | ···················· | |
| H | 6306 | hCoV-19/England/LEED-2AA103/2020\|EPI_ISL_538829\|20 | ···················· | ·A·················· | ····················· | ···················· | ···················· | ···················· | |
| H | 6307 | hCoV-19/England/LEED-2AA4EC/2020\|EPI_ISL_538830\|20 | ···················· | ···················· | ····················· | ···················· | ···················· | ···················· | |
| H | 6308 | hCoV-19/England/LEED-2A941A/2020\|EPI_ISL_538831\|20 | ···················· | ···················A | ····················· | ···················· | ···················· | ···················· | 1 |
| H | 6309 | hCoV-19/England/LEED-2AA4BF/2020\|EPI_ISL_538832\|20 | ···················· | ···················· | ····················· | ···················· | ···················· | ···················· | |
| H | 6310 | hCoV-19/England/LEED-2AA17C/2020\|EPI_ISL_538833\|20 | ···················· | ·R·················· | ····················· | ···················· | ···················· | ···················· | |
| H | 6311 | hCoV-19/England/LEED-2A9939/2020\|EPI_ISL_538834\|20 | ···················· | ···················· | ····················· | ···················· | ···················· | ···················· | |
| H | 6312 | hCoV-19/England/LEED-2A9A18/2020\|EPI_ISL_538835\|20 | ···················· | ·A·················· | ····················· | ···················· | ···················· | ···················· | |
| H | 6313 | hCoV-19/England/LEED-2AA0CA/2020\|EPI_ISL_538836\|20 | ···················· | ·A·················· | ····················· | ···················· | ···················· | ···················· | |
| H | 6314 | hCoV-19/England/LEED-2A9A63/2020\|EPI_ISL_538837\|20 | ···················· | ···················· | ····················· | ···················· | ···················· | ···················· | |
| H | 6315 | hCoV-19/England/LEED-2A9A72/2020\|EPI_ISL_538838\|20 | ···················· | ···················A | ····················· | ···················· | ···················· | ···················· | 1 |
| H | 6316 | hCoV-19/England/LEED-2A9C6D/2020\|EPI_ISL_538839\|20 | ···················· | ·A·················· | ····················· | ···················· | ···················· | ···················· | |
| H | 6317 | hCoV-19/England/LEED-2AA4DD/2020\|EPI_ISL_538840\|20 | ···················· | ···················· | ····················· | ···················· | ···················· | ···················· | |
| H | 6318 | hCoV-19/England/LEED-2AA14F/2020\|EPI_ISL_538842\|20 | ···················· | ···················· | ····················· | ···················· | ···················· | ···················· | |
| H | 6319 | hCoV-19/England/LEED-2AA31C/2020\|EPI_ISL_538843\|20 | ···················· | ·A·················· | ····················· | ···················· | ···················· | ···················· | |
| H | 6320 | hCoV-19/England/LEED-2AA2E2/2020\|EPI_ISL_538844\|20 | ···················· | ···················· | ····················· | ···················· | ···················· | ···················· | |
| H | 6321 | hCoV-19/England/LEED-2A9966/2020\|EPI_ISL_538845\|20 | ···················· | ···A················ | ····················· | ···················· | ···················· | ···················· | |
| H | 6322 | hCoV-19/England/LEED-2A9C21/2020\|EPI_ISL_538846\|20 | ···················· | ···················· | ····················· | ···················· | ···················· | ···················· | |
| H | 6323 | hCoV-19/England/LEED-2AA473/2020\|EPI_ISL_538847\|20 | ···················· | ···················· | ····················· | ···················· | ···················· | ···················· | |
| H | 6324 | hCoV-19/England/LEED-2A9429/2020\|EPI_ISL_538848\|20 | ···················· | ···················· | ····················· | ···················· | ···················· | ···················· | |
| H | 6325 | hCoV-19/England/LEED-2A9975/2020\|EPI_ISL_538849\|20 | ···················· | ···················· | ····················· | ···················· | ···················· | ···················· | |
| H | 6326 | hCoV-19/England/LEED-2AA437/2020\|EPI_ISL_538852\|20 | ···················· | ···················· | ····················· | ···················· | ···················· | ···················· | |
| H | 6327 | hCoV-19/England/LEED-2A9BBB/2020\|EPI_ISL_538853\|20 | ···················· | ···················· | ····················· | ···················· | ···················· | ···················· | |
| H | 6328 | hCoV-19/England/LEED-2AA0BB/2020\|EPI_ISL_538854\|20 | ···················· | ·A·················· | ····················· | ···················· | ···················· | ···················· | |
| H | 6329 | hCoV-19/England/LEED-2A9B51/2020\|EPI_ISL_538855\|20 | ···················· | ·R·················· | ····················· | ···················· | ···················· | ···················· | |
| H | 6330 | hCoV-19/England/LEED-2AA5E9/2020\|EPI_ISL_538856\|20 | ···················· | ···················· | ····················· | ···················· | ···················· | ···················· | |
| H | 6331 | hCoV-19/England/LEED-2AA349/2020\|EPI_ISL_538857\|20 | ···················· | ···················· | ····················· | ···················· | ···················· | ···················· | |
| H | 6332 | hCoV-19/England/LEED-2A9BAC/2020\|EPI_ISL_538858\|20 | ···················· | ···················· | ····················· | ···················· | ···················· | ···················· | |
| H | 6333 | hCoV-19/England/LEED-2AA67D/2020\|EPI_ISL_538859\|20 | ···················· | ···················· | ····················· | ···················· | ···················· | ···················· | |
| H | 6334 | hCoV-19/England/LEED-2A9B7F/2020\|EPI_ISL_538860\|20 | ···················· | ·A·················· | ····················· | ···················· | ···················· | ···················· | |
| H | 6335 | hCoV-19/England/LEED-2A9A45/2020\|EPI_ISL_538861\|20 | ···················· | ···················A | ····················· | ···················· | ···················· | ···················· | 1 |
| H | 6336 | hCoV-19/England/LEED-2A9957/2020\|EPI_ISL_538862\|20 | ···················· | ···················· | ····················· | ···················· | ···················· | ···················· | |
| H | 6337 | hCoV-19/England/LEED-2A94ED/2020\|EPI_ISL_538864\|20 | ···················· | ···················· | ····················· | ···················· | ···················· | ···················· | |
| H | 6338 | hCoV-19/England/LEED-2AA0D9/2020\|EPI_ISL_538865\|20 | ···················· | ···················· | ····················· | ···················· | ···················· | ···················· | |
| H | 6339 | hCoV-19/England/LEED-2A9AFA/2020\|EPI_ISL_538867\|20 | ···················· | ···················· | ····················· | ···················· | ···················· | ···················· | |
| H | 6340 | hCoV-19/England/LEED-2A98F0/2020\|EPI_ISL_538868\|20 | ···················· | ···················· | ····················· | ···················· | ···················· | ···················· | |
| H | 6341 | hCoV-19/England/LEED-2AA297/2020\|EPI_ISL_538869\|20 | ···················· | ···················· | ····················· | ···················· | ···················· | ···················· | |
| H | 6342 | hCoV-19/England/LEED-2AA051/2020\|EPI_ISL_538870\|20 | ···················· | ···················· | ····················· | ···················· | ···················· | ···················· | |
| H | 6343 | hCoV-19/England/LEED-2A985A/2020\|EPI_ISL_538871\|20 | ···················· | ···················· | ····················· | ···················· | ···················· | ···················· | |
| H | 6344 | hCoV-19/England/LEED-2A9447/2020\|EPI_ISL_538872\|20 | ···················· | ···················· | ····················· | ···················· | ···················· | ···················· | |
| H | 6345 | hCoV-19/England/LEED-2A9BE8/2020\|EPI_ISL_538874\|20 | ···················· | ···················· | ····················· | ···················· | ···················· | ···················· | |
| H | 6346 | hCoV-19/England/LEED-2A9878/2020\|EPI_ISL_538877\|20 | ···················· | ···················· | ····················· | ···················· | ···················· | ···················· | |
| H | 6347 | hCoV-19/England/LEED-2A992A/2020\|EPI_ISL_538878\|20 | ···················· | ···················· | ····················· | ···················· | ···················· | ···················· | |
| H | 6348 | hCoV-19/England/LEED-2A984B/2020\|EPI_ISL_538879\|20 | ···················· | ···················· | ····················· | ···················· | ···················· | ···················· | |
| H | 6349 | hCoV-19/England/LEED-2AA288/2020\|EPI_ISL_538880\|20 | ···················· | ·A·················· | ····················· | ···················· | ···················· | ···················· | |
| H | 6350 | hCoV-19/England/LEED-2AA121/2020\|EPI_ISL_538881\|20 | ···················· | ···················· | ····················· | ···················· | ···················· | ···················· | |
| H | 6351 | hCoV-19/England/LEED-2AA59E/2020\|EPI_ISL_538882\|20 | ···················· | ···················· | ····················· | ···················· | ···················· | ···················· | |
| H | 6352 | hCoV-19/England/LEED-2A9438/2020\|EPI_ISL_538883\|20 | ···················· | ···················· | ····················· | ···················· | ···················· | ···················· | |
| H | 6353 | hCoV-19/England/LEED-2A9A54/2020\|EPI_ISL_538885\|20 | ···················· | ···················· | ····················· | ···················· | ···················· | ···················· | |
| H | 6354 | hCoV-19/England/LEED-2AA3EF/2020\|EPI_ISL_538886\|20 | ···················· | ···················· | ····················· | ···················· | ···················· | ···················· | |
| H | 6355 | hCoV-19/England/LEED-2A9896/2020\|EPI_ISL_538887\|20 | ···················· | ···················· | ····················· | ···················· | ···················· | ···················· | |
| H | 6356 | hCoV-19/England/LEED-2A9508/2020\|EPI_ISL_538888\|20 | ···················· | ···················· | ····················· | ···················· | ···················· | ···················· | |
| H | 6357 | hCoV-19/England/LEED-2AA09D/2020\|EPI_ISL_538889\|20 | ···················· | ···················· | ····················· | ···················· | ···················· | ···················· | |

| | | | | | | | | |
|---|---|---|---|---|---|---|---|---|
| H | 6358 hCoV-19/USA/NMDOH-2020321648/2020\|EPI_ISL_542070\|2 | ···················· | ···················· | ···················· | ···················· | ···················· | ···················· | |
| H | 6359 hCoV-19/USA/NMDOH-2020321813/2020\|EPI_ISL_542071\|2 | ···················· | ···················· | ···················· | ···················· | ···················· | ···················· | |
| H | 6360 hCoV-19/USA/NMDOH-2020321832/2020\|EPI_ISL_542072\|2 | ···················· | ·A·················· | ···················· | ···················· | ···················· | ···················· | |
| H | 6361 hCoV-19/USA/NMDOH-2020321845/2020\|EPI_ISL_542073\|2 | ···················· | ·A·················· | ···················· | ···················· | ···················· | ···················· | |
| H | 6362 hCoV-19/USA/NMDOH-2020321846/2020\|EPI_ISL_542074\|2 | ···················· | ·A·················· | ···················· | ···················· | ···················· | ···················· | |
| H | 6363 hCoV-19/USA/NMDOH-2020322105/2020\|EPI_ISL_542077\|2 | ···················· | ···················· | ···················· | ···················· | ···················· | ···················· | |
| H | 6364 hCoV-19/USA/NMDOH-2020322123/2020\|EPI_ISL_542078\|2 | ···················· | ···················· | ···················· | ···················· | ···················· | ···················· | |
| H | 6365 hCoV-19/USA/NMDOH-2020322124/2020\|EPI_ISL_542079\|2 | ···················· | ···················· | ···················· | ···················· | ···················· | ···················· | |
| H | 6366 hCoV-19/USA/NMDOH-2020322125/2020\|EPI_ISL_542080\|2 | ···················· | ···················· | ···················· | ···················· | ···················· | ···················· | |
| H | 6367 hCoV-19/USA/NMDOH-2020322173/2020\|EPI_ISL_542082\|2 | ···················· | ···················· | ···················· | ···················· | ···················· | ···················· | |
| H | 6368 hCoV-19/USA/NMDOH-2020322270/2020\|EPI_ISL_542083\|2 | ···················· | ···················· | ···················· | ···················· | ···················· | ···················· | |
| H | 6369 hCoV-19/USA/NMDOH-2020322355/2020\|EPI_ISL_542084\|2 | ···················· | ···················· | ···················· | ···················· | ···················· | ···················· | |
| H | 6370 hCoV-19/USA/NMDOH-2020323484/2020\|EPI_ISL_542086\|2 | ···················· | ···················· | ···················· | ···················· | ···················· | ···················· | |
| H | 6371 hCoV-19/USA/NMDOH-2020323515/2020\|EPI_ISL_542087\|2 | ···················· | ···················· | ···················· | ···················· | ···················· | ···················· | |
| H | 6372 hCoV-19/USA/NMDOH-2020323670/2020\|EPI_ISL_542088\|2 | ···················· | ···················· | ···················· | ···················· | ···················· | ···················· | |
| H | 6373 hCoV-19/USA/NMDOH-2020323679/2020\|EPI_ISL_542089\|2 | ···················· | ···················· | ···················· | ···················· | ···················· | ···················· | |
| H | 6374 hCoV-19/USA/NMDOH-2020323686/2020\|EPI_ISL_542090\|2 | ···················· | ···················· | ···················· | ···················· | ···················· | ···················· | |
| H | 6375 hCoV-19/USA/NMDOH-2020324000/2020\|EPI_ISL_542091\|2 | ···················· | ···················· | ···················· | ···················· | ···················· | ···················· | |
| H | 6376 hCoV-19/USA/NMDOH-2020324045/2020\|EPI_ISL_542092\|2 | ···················· | ···················· | ···················· | ···················· | ···················· | ···················· | |
| H | 6377 hCoV-19/USA/NMDOH-2020324052/2020\|EPI_ISL_542093\|2 | ···················· | ····A··············· | ···················· | ···················· | ···················· | ···················· | |
| H | 6378 hCoV-19/USA/NMDOH-2020324233/2020\|EPI_ISL_542094\|2 | ···················· | ···················· | ···················· | ···················· | ···················· | ···················· | |
| H | 6379 hCoV-19/USA/NMDOH-2020324359/2020\|EPI_ISL_542095\|2 | ···················· | ·A·················· | ···················· | ···················· | ···················· | ···················· | |
| H | 6380 hCoV-19/USA/NMDOH-2020325588/2020\|EPI_ISL_542097\|2 | ···················· | ···················· | ···················· | ···················· | ···················· | ···················· | |
| H | 6381 hCoV-19/Italy/LOM-UniMI-L100/2020\|EPI_ISL_542098\|2 | ···················· | ···················· | ···················· | ···················· | ···················· | ···················· | |
| H | 6382 hCoV-19/Italy/LOM-UniMI-L101/2020\|EPI_ISL_542099\|2 | ···················· | ···················· | ···················· | ···················· | ···················· | ···················· | |
| H | 6383 hCoV-19/USA/TX-HMH-4034/2020\|EPI_ISL_543314\|2020-0 | ···················· | ···················· | ···················· | ···················· | ···················· | ···················· | |
| H | 6385 hCoV-19/USA/TX-HMH-4040/2020\|EPI_ISL_543318\|2020-0 | ···················· | ···················· | ···················· | ···················· | ···················· | ···················· | |
| H | 6386 hCoV-19/USA/TX-HMH-4041/2020\|EPI_ISL_543319\|2020-0 | ···················· | ···················· | ···················· | ···················· | ···················· | ···················· | |
| H | 6387 hCoV-19/USA/TX-HMH-4042/2020\|EPI_ISL_543320\|2020-0 | ···················· | ···················· | ···················· | ···················· | ···················· | ···················· | |
| H | 6388 hCoV-19/USA/TX-HMH-4044/2020\|EPI_ISL_543321\|2020-0 | ···················· | ···················· | ···················· | ···················· | ···················· | ···················· | |
| H | 6389 hCoV-19/USA/TX-HMH-4045/2020\|EPI_ISL_543322\|2020-0 | ···················· | ···················· | ···················· | ···················· | ···················· | ···················· | |
| H | 6390 hCoV-19/USA/TX-HMH-4046/2020\|EPI_ISL_543323\|2020-0 | ···················· | ···················· | ···················· | ···················· | ···················· | ···················· | |
| H | 6391 hCoV-19/USA/TX-HMH-4047/2020\|EPI_ISL_543324\|2020-0 | ···················· | ···················· | ···················· | ···················· | ···················· | ···················· | |
| H | 6392 hCoV-19/USA/TX-HMH-4048/2020\|EPI_ISL_543325\|2020-0 | ···················· | ···················· | ···················· | ···················· | ···················· | ···················· | |
| H | 6393 hCoV-19/USA/TX-HMH-4050/2020\|EPI_ISL_543326\|2020-0 | ···················· | ···················· | ···················· | ···················· | ···················· | ···················· | |
| H | 6395 hCoV-19/USA/TX-HMH-4052/2020\|EPI_ISL_543328\|2020-0 | ···················· | ···················· | ···················· | ···················· | ···················· | ···················· | |
| H | 6396 hCoV-19/USA/TX-HMH-4053/2020\|EPI_ISL_543329\|2020-0 | ···················· | ···················· | ···················· | ···················· | ···················· | ···················· | |
| H | 6397 hCoV-19/USA/TX-HMH-4055/2020\|EPI_ISL_543331\|2020-0 | ···················· | ···················· | ···················· | ···················· | ··············N····· | NN·N················ | 1 |
| H | 6398 hCoV-19/USA/TX-HMH-4056/2020\|EPI_ISL_543332\|2020-0 | ···················· | ···················· | ···················· | ···················· | ···················· | ···················· | |
| H | 6399 hCoV-19/USA/TX-HMH-4060/2020\|EPI_ISL_543334\|2020-0 | ···················· | ·············A······ | ···················· | ···················· | ··············NN···· | NN·················· | |
| H | 6400 hCoV-19/USA/TX-HMH-4061/2020\|EPI_ISL_543335\|2020-0 | ···················· | ···················· | ···················· | ···················· | ···················· | ···················· | |
| H | 6401 hCoV-19/Northern Ireland/QEUH-96ABDD/2020\|EPI_ISL_ | ···················· | ···················· | ···················· | ···················· | ···················· | ···················· | |
| H | 6402 hCoV-19/England/QEUH-96AC9E/2020\|EPI_ISL_531359\|20 | ···················· | ···················· | ···················· | ···················· | ···················· | ···················· | |
| H | 6403 hCoV-19/Northern Ireland/QEUH-96B83F/2020\|EPI_ISL_ | ···················· | ···················· | ···················· | ···················· | ···················· | ···················· | |
| H | 6404 hCoV-19/England/QEUH-96BF76/2020\|EPI_ISL_531362\|20 | ···················· | ···················· | ···················· | ···················· | ···················· | ···················· | |
| H | 6405 hCoV-19/England/QEUH-96B170/2020\|EPI_ISL_531363\|20 | ···················· | ···················· | ···················· | ···················· | ···················· | ···················· | |
| H | 6406 hCoV-19/England/QEUH-96BE88/2020\|EPI_ISL_531364\|20 | ···················· | ···················· | ···················· | ···················· | ···················· | ···················· | |
| H | 6407 hCoV-19/Scotland/QEUH-96B5A1/2020\|EPI_ISL_531365\|2 | ···················· | ···················· | ···················· | ···················· | ···················· | ···················· | |
| H | 6408 hCoV-19/England/QEUH-96AEB6/2020\|EPI_ISL_531366\|20 | ···················· | ···················· | ···················· | ···················· | ···················· | ···················· | |
| H | 6409 hCoV-19/England/QEUH-96B50B/2020\|EPI_ISL_531367\|20 | ···················· | ···················· | ···················· | ···················· | ···················· | ···················· | |
| H | 6410 hCoV-19/England/QEUH-96B5FC/2020\|EPI_ISL_531368\|20 | ···················· | ···················· | ···················· | ···················· | ···················· | ···················· | |
| H | 6411 hCoV-19/England/QEUH-96B8B7/2020\|EPI_ISL_531369\|20 | ···················· | ···················· | ···················· | ···················· | ···················· | ···················· | |
| H | 6412 hCoV-19/England/QEUH-96B0A0/2020\|EPI_ISL_531370\|20 | ···················· | ·A·················· | ···················· | ···················· | ···················· | ···················· | |
| H | 6413 hCoV-19/England/QEUH-96BD03/2020\|EPI_ISL_531371\|20 | ···················· | ···················· | ···················· | ···················· | ···················· | ···················· | |
| H | 6414 hCoV-19/England/QEUH-96AC8F/2020\|EPI_ISL_531372\|20 | ···················· | ·A·················· | ···················· | ···················· | ···················· | ···················· | |
| H | 6415 hCoV-19/England/QEUH-96BEE2/2020\|EPI_ISL_531373\|20 | ···················· | ···················· | ···················· | ···················· | ···················· | ···················· | |
| H | 6416 hCoV-19/Scotland/QEUH-96B4E0/2020\|EPI_ISL_531374\|2 | ···················· | ···················· | ···················· | ···················· | ···················· | ···················· | |
| H | 6417 hCoV-19/Wales/QEUH-96B820/2020\|EPI_ISL_531375\|2020 | ···················· | ···················· | ···················· | ···················· | ···················· | ···················· | |
| H | 6418 hCoV-19/England/QEUH-96B019/2020\|EPI_ISL_531376\|20 | ···················· | ···················· | ···················· | ···················· | ···················· | ···················· | |
| H | 6419 hCoV-19/England/QEUH-96ADC8/2020\|EPI_ISL_531377\|20 | ···················· | ···················· | ···················· | ···················· | ···················· | ···················· | |
| H | 6420 hCoV-19/Northern Ireland/QEUH-96A9F1/2020\|EPI_ISL_ | ···················· | ···················· | ···················· | ···················· | ···················· | ···················· | |
| H | 6421 hCoV-19/England/QEUH-96B4C2/2020\|EPI_ISL_531380\|20 | ···················· | ···················· | ···················· | ···················· | ···················· | ···················· | |
| H | 6422 hCoV-19/Scotland/QEUH-96A95B/2020\|EPI_ISL_531382\|2 | ···················· | ···················· | ···················· | ···················· | ···················· | ···················· | |
| H | 6423 hCoV-19/England/QEUH-96AF3B/2020\|EPI_ISL_531383\|20 | ···················· | ···················· | ···················· | ···················· | ····T··············· | ···················· | |
| H | 6424 hCoV-19/England/QEUH-96B2B9/2020\|EPI_ISL_531385\|20 | ···················· | ···················· | ···················· | ···················· | ···················· | ···················· | |
| H | 6425 hCoV-19/Northern Ireland/QEUH-96B8D5/2020\|EPI_ISL_ | ···················· | ···················· | ···················· | ···················· | ···················· | ···················· | |
| H | 6426 hCoV-19/England/QEUH-96AD9B/2020\|EPI_ISL_531389\|20 | ···················· | ···················· | ···················· | ···················· | ···················· | ···················· | |
| H | 6427 hCoV-19/England/QEUH-96BE5B/2020\|EPI_ISL_531390\|20 | ···················· | ·A·······A·········· | ···················· | ···················· | ···················· | ···················· | |
| H | 6428 hCoV-19/England/QEUH-96AF0E/2020\|EPI_ISL_531392\|20 | ···················· | ·A·················· | ···················· | ···················· | ···················· | ···················· | |
| H | 6429 hCoV-19/Northern Ireland/QEUH-96A9D3/2020\|EPI_ISL_ | ···················· | ···················· | ···················· | ···················· | ······T············· | ···················· | |
| H | 6430 hCoV-19/England/QEUH-96B222/2020\|EPI_ISL_531394\|20 | ···················· | ·A·················· | ···················· | ···················· | ···················· | ···················· | |
| H | 6431 hCoV-19/England/QEUH-96B3E3/2020\|EPI_ISL_531396\|20 | ···················· | ···················· | ···················· | ···················· | ···················· | ···················· | |
| H | 6432 hCoV-19/England/QEUH-96BC42/2020\|EPI_ISL_531397\|20 | ···T················ | ···················· | ···················· | ···················· | ···················· | ···················· | |
| H | 6433 hCoV-19/Northern Ireland/QEUH-96A94C/2020\|EPI_ISL_ | ···················· | ···················· | ···················· | ···················· | ···················· | ···················· | |
| H | 6434 hCoV-19/England/QEUH-96BEF1/2020\|EPI_ISL_531399\|20 | ···················· | ···················· | ···················· | ···················· | ···················· | ···················· | |
| H | 6435 hCoV-19/England/QEUH-96AD7D/2020\|EPI_ISL_531400\|20 | ···················· | ···················· | ···················· | ···················· | ···················· | ···················· | |
| H | 6436 hCoV-19/England/QEUH-96BDC7/2020\|EPI_ISL_531401\|20 | ···················· | ···················· | ···················· | ···················· | ···················· | ···················· | |
| H | 6437 hCoV-19/England/QEUH-96BAFD/2020\|EPI_ISL_531402\|20 | ···················· | ·A·················· | ···················· | ···················· | ···················· | ···A················ | |
| H | 6438 hCoV-19/England/QEUH-96B88A/2020\|EPI_ISL_531403\|20 | ···················· | ···················· | ···················· | ···················· | ···················· | ···················· | |
| H | 6439 hCoV-19/Wales/QEUH-96BF85/2020\|EPI_ISL_531404\|2020 | ···················· | ···················· | ···················· | ···················· | ···················· | ···················· | |

| | | | | | | | | |
|---|---|---|---|---|---|---|---|---|
| H | 6440 hCoV-19/Northern Ireland/QEUH-96A92E/2020\|EPI_ISL_ | ···················· | ···················· | ···················· | ···················· | ···················· | ···················· | |
| H | 6441 hCoV-19/England/QEUH-96B1AD/2020\|EPI_ISL_531407\|20 | ···················· | ···················· | ···················· | ···················· | ···················· | ···················· | |
| H | 6442 hCoV-19/England/QEUH-96BB36/2020\|EPI_ISL_531408\|20 | ···················· | ···················· | ···················· | ···················· | ···················· | ···················· | |
| H | 6443 hCoV-19/Northern Ireland/QEUH-96AF95/2020\|EPI_ISL_ | ···················· | ···················· | ···················· | ···················· | ···················· | ···················· | |
| H | 6444 hCoV-19/England/QEUH-96AFA4/2020\|EPI_ISL_531410\|20 | ···················· | ···················· | ···················· | ···················· | ···················· | ···················· | |
| H | 6445 hCoV-19/England/QEUH-96B6F9/2020\|EPI_ISL_531411\|20 | ···················· | ···················· | ···················· | ···················· | ··················C· | ···················· | 1 |
| H | 6446 hCoV-19/Scotland/QEUH-96AE6B/2020\|EPI_ISL_531412\|2 | ···················· | ···················· | ···················· | ···················· | ···················· | ···················· | |
| H | 6447 hCoV-19/England/QEUH-96BA84/2020\|EPI_ISL_531413\|20 | ···················· | ···················· | ···················· | ···················· | ···················· | ···················· | |
| H | 6448 hCoV-19/England/QEUH-96B592/2020\|EPI_ISL_531414\|20 | ···················· | ···················· | ···················· | ···················· | ···················· | ···················· | |
| H | 6449 hCoV-19/England/QEUH-96B0BF/2020\|EPI_ISL_531415\|20 | ···················· | ···················· | ···················· | ···················· | ···················· | ···················· | |
| H | 6450 hCoV-19/Scotland/QEUH-96B28C/2020\|EPI_ISL_531416\|2 | ···················· | ···················· | ···················· | ···················· | ···················· | ···················· | |
| H | 6451 hCoV-19/Northern Ireland/QEUH-96B7E7/2020\|EPI_ISL_ | ···················· | ···················· | ···················· | ···················· | ···················· | ···················· | |
| H | 6452 hCoV-19/Northern Ireland/QEUH-96AAA3/2020\|EPI_ISL_ | ···················· | ···················· | ···················· | ···················· | ···················· | ···················· | |
| H | 6453 hCoV-19/England/QEUH-96BE4C/2020\|EPI_ISL_531419\|20 | ···················· | ·A······A··········· | ···················· | ···················· | ···················· | ···················· | |
| H | 6454 hCoV-19/Scotland/QEUH-96B27D/2020\|EPI_ISL_531420\|2 | ···················· | ···················· | ···················· | ···················· | ···················· | ···················· | |
| H | 6455 hCoV-19/Scotland/QEUH-96B680/2020\|EPI_ISL_531422\|2 | ···················· | ···················· | ···················· | ···················· | ···················· | ···················· | |
| H | 6456 hCoV-19/Northern Ireland/QEUH-96B811/2020\|EPI_ISL_ | ···················· | ···················· | ···················· | ···················· | ···················· | ···················· | |
| H | 6457 hCoV-19/Northern Ireland/QEUH-96ABBF/2020\|EPI_ISL_ | ···················· | ···················· | ···················· | ···················· | ···················· | ···················· | |
| H | 6458 hCoV-19/Northern Ireland/QEUH-96B93C/2020\|EPI_ISL_ | ···················· | ···················· | ···················· | ···················· | ···················· | ···················· | |
| H | 6459 hCoV-19/England/QEUH-96BC9D/2020\|EPI_ISL_531427\|20 | ···················· | ···················· | ···················· | ···················· | ···················· | ···················· | |
| H | 6460 hCoV-19/Northern Ireland/QEUH-96AAB2/2020\|EPI_ISL_ | ···················· | ···················· | ···················· | ···················· | ···················· | ···················· | |
| H | 6461 hCoV-19/England/QEUH-96BA93/2020\|EPI_ISL_531429\|20 | ···················· | ···················· | ···················· | ···················· | ···················· | ···················· | |
| H | 6462 hCoV-19/England/QEUH-96BB72/2020\|EPI_ISL_531431\|20 | ···················· | ···················· | ···················· | ···················· | ···················· | ···················· | |
| H | 6463 hCoV-19/Scotland/QEUH-96B161/2020\|EPI_ISL_531432\|2 | ···················· | ···················· | ···················· | ···················· | ···················· | ···················· | |
| H | 6464 hCoV-19/Wales/QEUH-96BF94/2020\|EPI_ISL_531433\|2020 | ···················· | ···················· | ···················· | ···················· | ···················· | ···················· | |
| H | 6465 hCoV-19/England/QEUH-96B486/2020\|EPI_ISL_531434\|20 | ···················· | ···················· | ···················· | ···················· | ···················· | ···················· | |
| H | 6466 hCoV-19/England/QEUH-96AC70/2020\|EPI_ISL_531435\|20 | ···················· | ···················· | ···················· | ···················· | ···················· | ···················· | |
| H | 6467 hCoV-19/England/QEUH-96AD31/2020\|EPI_ISL_531436\|20 | ···················· | ···················· | ···················· | ···················· | ···················· | ···················· | |
| H | 6468 hCoV-19/England/QEUH-96B565/2020\|EPI_ISL_531437\|20 | ···················· | ···················· | ···················· | ···················· | ···················· | ···················· | |
| H | 6469 hCoV-19/England/QEUH-96B750/2020\|EPI_ISL_531439\|20 | ···················· | ···················· | ···················· | ···················· | ·····T·············· | ···················· | |
| H | 6470 hCoV-19/England/QEUH-89C2DE/2020\|EPI_ISL_532688\|20 | ···················· | ···················· | ···················· | ···················· | ···················· | ···················· | |
| H | 6471 hCoV-19/England/QEUH-88CEFB/2020\|EPI_ISL_532690\|20 | ···················· | ···················· | ···················· | ···················· | ···················· | ···················· | |
| H | 6472 hCoV-19/England/QEUH-880E33/2020\|EPI_ISL_532691\|20 | ···················· | ···················· | ···················· | ···················· | ···················· | ···················· | |
| H | 6473 hCoV-19/England/QEUH-8A08E4/2020\|EPI_ISL_532692\|20 | ···················· | ···················· | ···················· | ···················· | ···················· | ···················· | |
| H | 6474 hCoV-19/Wales/QEUH-8ADD60/2020\|EPI_ISL_532693\|2020 | ···················· | ···················· | ···················· | ···················· | ···················· | ···················· | |
| H | 6475 hCoV-19/Northern Ireland/QEUH-88B287/2020\|EPI_ISL_ | ···················· | ···················· | ···················· | ···················· | ···················· | ···················· | |
| H | 6476 hCoV-19/England/QEUH-8979B0/2020\|EPI_ISL_532695\|20 | ···················· | ···················· | ···················· | ···················· | ···················· | ···················· | |
| H | 6477 hCoV-19/Northern Ireland/QEUH-8806E0/2020\|EPI_ISL_ | ···················· | ···················· | ···················· | ···················· | ···················· | ···················· | |
| H | 6478 hCoV-19/Scotland/QEUH-89DB2D/2020\|EPI_ISL_532697\|2 | ···················· | ···················· | ···················· | ···················· | ···················· | ···················· | |
| H | 6479 hCoV-19/Northern Ireland/QEUH-88ED65/2020\|EPI_ISL_ | ···················· | ···················· | ···················· | ···················· | ···················· | ···················· | |
| H | 6480 hCoV-19/England/QEUH-88C7A5/2020\|EPI_ISL_532699\|20 | ···················· | ···················· | ···················· | ···················· | ···················· | ···················· | |
| H | 6481 hCoV-19/England/QEUH-881EC9/2020\|EPI_ISL_532700\|20 | ···················· | ···················· | ···················· | ···················· | ···················· | ···················· | |
| H | 6482 hCoV-19/England/QEUH-8AF5DF/2020\|EPI_ISL_532701\|20 | ···················· | ···················· | ···················· | ···················· | ···················· | ···················· | |
| H | 6483 hCoV-19/England/QEUH-894133/2020\|EPI_ISL_532702\|20 | ···················· | ···················· | ···················· | ···················· | ···················· | ···················· | |
| H | 6484 hCoV-19/England/QEUH-894C9C/2020\|EPI_ISL_532703\|20 | ···················· | ···················· | ···················· | ···················· | ···················· | ···················· | |
| H | 6485 hCoV-19/England/QEUH-89531E/2020\|EPI_ISL_532704\|20 | ···················· | ···················· | ···················· | ···················· | ···················· | ···················· | |
| H | 6486 hCoV-19/Scotland/QEUH-87C2F2/2020\|EPI_ISL_532705\|2 | ···················· | ···················· | ···················· | ···················· | ···················· | ···················· | |
| H | 6487 hCoV-19/England/QEUH-87FF70/2020\|EPI_ISL_532706\|20 | ···················· | ···················· | ···················· | ···················· | ···················· | ···················· | |
| H | 6488 hCoV-19/England/QEUH-8865ED/2020\|EPI_ISL_532708\|20 | ···················· | ···················· | ···················· | ···················· | ···················· | ···················· | |
| H | 6489 hCoV-19/England/QEUH-8AF029/2020\|EPI_ISL_532711\|20 | ···················· | ···················· | ···················· | ···················· | ···················· | ···················· | |
| H | 6490 hCoV-19/England/QEUH-8B3909/2020\|EPI_ISL_532712\|20 | ···················· | ···················· | ···················· | ···················· | ···················· | ···················· | |
| H | 6491 hCoV-19/England/QEUH-89AF24/2020\|EPI_ISL_532713\|20 | ···················· | ···················· | ···················· | ···················· | ···················· | ···················· | |
| H | 6492 hCoV-19/England/QEUH-881342/2020\|EPI_ISL_532714\|20 | ···················· | ···················· | ···················· | ···················· | ···················· | ···················· | |
| H | 6493 hCoV-19/Scotland/QEUH-87EF35/2020\|EPI_ISL_532715\|2 | ···················· | ···················· | ···················· | ···················· | ···················· | ···················· | |
| H | 6494 hCoV-19/England/QEUH-8AE7E9/2020\|EPI_ISL_532716\|20 | ···················· | ···················· | ···················· | ···················· | ···················· | ···················· | |
| H | 6495 hCoV-19/England/QEUH-8AE576/2020\|EPI_ISL_532717\|20 | ···················· | ···················· | ···················· | ···················· | ···················· | ···················· | |
| H | 6496 hCoV-19/England/QEUH-886D12/2020\|EPI_ISL_532718\|20 | ···················· | ···················· | ···················· | ···················· | ···················· | ···················· | |
| H | 6497 hCoV-19/England/QEUH-8AE3B8/2020\|EPI_ISL_532719\|20 | ···················· | ···················· | ···················· | ···················· | ···················· | ···················· | |
| H | 6498 hCoV-19/England/QEUH-8AD331/2020\|EPI_ISL_532721\|20 | ···················· | ···················· | ···················· | ···················· | ···················· | ···················· | |
| H | 6499 hCoV-19/England/QEUH-885FE0/2020\|EPI_ISL_532722\|20 | ···················· | ···················· | ···················· | ···················· | ···················· | ···················· | |
| H | 6500 hCoV-19/Northern Ireland/QEUH-8AB3D9/2020\|EPI_ISL_ | ···················· | ···················· | ···················· | ···················· | ···················· | ···················· | |
| H | 6501 hCoV-19/England/QEUH-87FC6A/2020\|EPI_ISL_532725\|20 | ···················· | ···················· | ···················· | ···················· | ···················· | ···················· | |
| H | 6502 hCoV-19/England/QEUH-8845FE/2020\|EPI_ISL_532726\|20 | ···················· | ···················· | ···················· | ···················· | ···················· | ···················· | |
| H | 6503 hCoV-19/England/QEUH-8A7D1B/2020\|EPI_ISL_532727\|20 | ···················· | ···················· | ···················· | ···················· | ···················· | ···················· | |
| H | 6504 hCoV-19/England/QEUH-89E2AF/2020\|EPI_ISL_532729\|20 | ···················· | ···················· | ···················· | ···················· | ···················· | ···················· | |
| H | 6505 hCoV-19/England/QEUH-89EA01/2020\|EPI_ISL_532730\|20 | ···················· | ···················· | ···················· | ···················· | ···················· | ···················· | |
| H | 6506 hCoV-19/England/QEUH-89523F/2020\|EPI_ISL_532731\|20 | ···················· | ···················· | ···················· | ···················· | ···················· | ···················· | |
| H | 6507 hCoV-19/England/QEUH-87D4CE/2020\|EPI_ISL_532732\|20 | ···················· | ·A·················· | ···················· | ···················· | ···················· | ···················· | |
| H | 6508 hCoV-19/Scotland/QEUH-89E46D/2020\|EPI_ISL_532733\|2 | ···················· | ···················· | ···················· | ···················· | ···················· | ···················· | |
| H | 6509 hCoV-19/England/QEUH-8988EF/2020\|EPI_ISL_532734\|20 | ···················· | ···················· | ···················· | ···················· | ···················· | ···················· | |
| H | 6510 hCoV-19/England/QEUH-89AE36/2020\|EPI_ISL_532735\|20 | ···················· | ···················· | ···················· | ···················· | ···················· | ···················· | |
| H | 6511 hCoV-19/England/QEUH-897A35/2020\|EPI_ISL_532736\|20 | ···················· | ···················· | ···················· | ···················· | ···················· | ···················· | |
| H | 6512 hCoV-19/England/QEUH-884804/2020\|EPI_ISL_532737\|20 | ···················· | ···················· | ···················· | ···················· | ···················· | ···················· | |
| H | 6513 hCoV-19/Scotland/QEUH-89E6D0/2020\|EPI_ISL_532738\|2 | ···················· | ···················· | ···················· | ···················· | ···················· | ···················· | |
| H | 6514 hCoV-19/England/QEUH-87F15C/2020\|EPI_ISL_532740\|20 | ···················· | ···················· | ···················· | ···················· | ···················· | ···················· | |
| H | 6515 hCoV-19/Northern Ireland/QEUH-88E8CE/2020\|EPI_ISL_ | ···················· | ···················· | ···················· | ···················· | ···················· | ···················· | |
| H | 6516 hCoV-19/England/QEUH-8953B4/2020\|EPI_ISL_532742\|20 | ···················· | ···················· | ···················· | ···················· | ···················· | ···················· | |
| H | 6517 hCoV-19/England/QEUH-88C8C0/2020\|EPI_ISL_532743\|20 | ···················· | ·A·················· | ···················· | ···················· | ···················· | ···················· | |
| H | 6518 hCoV-19/Scotland/QEUH-895967/2020\|EPI_ISL_532744\|2 | ···················· | ···················· | ···················· | ···················· | ···················· | ···················· | |
| H | 6519 hCoV-19/England/QEUH-883C81/2020\|EPI_ISL_532745\|20 | ···················· | ···················· | ···················· | ···················· | ···················· | ···················· | |

| | | | | | | | |
|---|---|---|---|---|---|---|---|
| H | 6520 hCoV-19/Scotland/QEUH-88C08C/2020\|EPI_ISL_532746\|2 | ···················· | ···················· | ···················· | ···················· | ···················· | ···················· |
| H | 6521 hCoV-19/England/QEUH-8A0AB1/2020\|EPI_ISL_532747\|20 | ···················· | ···················· | ···················· | ···················· | ···················· | ···················· |
| H | 6522 hCoV-19/England/QEUH-889CA9/2020\|EPI_ISL_532748\|20 | ···················· | ···················· | ···················· | ···················· | ···················· | ···················· |
| H | 6523 hCoV-19/England/QEUH-8A02F5/2020\|EPI_ISL_532749\|20 | ···················· | ···················· | ···················· | ···················· | ···················· | ···················· |
| H | 6524 hCoV-19/Scotland/QEUH-887DB7/2020\|EPI_ISL_532750\|2 | ···················· | ···················· | ···················· | ···················· | ···················· | ···················· |
| H | 6525 hCoV-19/England/QEUH-8997A5/2020\|EPI_ISL_532751\|20 | ···················· | ···················· | ···················· | ···················· | ···················· | ···················· |
| H | 6526 hCoV-19/England/QEUH-8854C3/2020\|EPI_ISL_532752\|20 | ···················· | ···················· | ···················· | ···················· | ···················· | ···················· |
| H | 6527 hCoV-19/Scotland/QEUH-8937D8/2020\|EPI_ISL_532753\|2 | ···················· | ·A·················· | ···················· | ···················· | ···················· | ···A················ |
| H | 6528 hCoV-19/England/QEUH-894AED/2020\|EPI_ISL_532754\|20 | ···················· | ···················· | ···················· | ···················· | ···················· | ···················· |
| H | 6529 hCoV-19/Scotland/QEUH-899FDA/2020\|EPI_ISL_532755\|2 | ···················· | ···················· | ···················· | ···················· | ···················· | ···················· |
| H | 6530 hCoV-19/England/QEUH-8819C8/2020\|EPI_ISL_532756\|20 | ···················· | ···················· | ···················· | ···················· | ···················· | ···················· |
| H | 6531 hCoV-19/Northern Ireland/QEUH-88255A/2020\|EPI_ISL_ | ···················· | ···················· | ···················· | ···················· | ···················· | ···················· |
| H | 6532 hCoV-19/Northern Ireland/QEUH-88F0D4/2020\|EPI_ISL_ | ···················· | ···················· | ···················· | ···················· | ···················· | ···················· |
| H | 6533 hCoV-19/England/QEUH-87DD1E/2020\|EPI_ISL_532760\|20 | ···················· | ···················· | ···················· | ···················· | ···················· | ···················· |
| H | 6534 hCoV-19/Wales/QEUH-8AED32/2020\|EPI_ISL_532761\|2020 | ···················· | ···················· | ···················· | ···················· | ···················· | ···················· |
| H | 6535 hCoV-19/England/QEUH-898560/2020\|EPI_ISL_532762\|20 | ···················· | ···················· | ···················· | ···················· | ···················· | ···················· |
| H | 6536 hCoV-19/Northern Ireland/QEUH-8AC9A9/2020\|EPI_ISL_ | ···················· | ···················· | ···················· | ···················· | ···················· | ···················· |
| H | 6537 hCoV-19/England/QEUH-8928F4/2020\|EPI_ISL_532764\|20 | ···················· | ···················· | ···················· | ···················· | ···················· | ···················· |
| H | 6538 hCoV-19/England/QEUH-89966C/2020\|EPI_ISL_532766\|20 | ···················· | ···················· | ···················· | ···················· | ···················· | ···················· |
| H | 6539 hCoV-19/Scotland/QEUH-89DD18/2020\|EPI_ISL_532767\|2 | ···················· | ···················· | ···················· | ···················· | ···················· | ···················· |
| H | 6540 hCoV-19/Northern Ireland/QEUH-8AB333/2020\|EPI_ISL_ | ···················· | ···················· | ···················· | ···················· | ···················· | ···················· |
| H | 6541 hCoV-19/England/QEUH-8A00BF/2020\|EPI_ISL_532769\|20 | ···················· | ···················· | ···················· | ···················· | ···················· | ···················· |
| H | 6542 hCoV-19/England/QEUH-8A971B/2020\|EPI_ISL_532771\|20 | ···················· | ···················· | ···················· | ···················· | ···················· | ···················· |
| H | 6543 hCoV-19/England/LEED-2A9465/2020\|EPI_ISL_538800\|20 | ···················· | ···················· | ···················· | ···················· | ···················· | ···················· |
| H | 6544 hCoV-19/England/LEED-2A9D00/2020\|EPI_ISL_538802\|20 | ···················· | ···················· | ···················· | ···················· | ···················· | ···················· |
| H | 6545 hCoV-19/England/LEED-2A9887/2020\|EPI_ISL_538803\|20 | ···················· | ···················· | ···················· | ···················· | ···················· | ···················· |
| H | 6546 hCoV-19/USA/NMDOH-2020117406/2020\|EPI_ISL_542014\|2 | ···················· | ···················· | ···················· | ···················· | ···················· | ···················· |
| H | 6547 hCoV-19/USA/NMDOH-2020117650/2020\|EPI_ISL_542016\|2 | ···················· | ···················· | ···················· | ···················· | ···················· | ···················· |
| H | 6548 hCoV-19/USA/NMDOH-2020117722/2020\|EPI_ISL_542017\|2 | ···················· | ···················· | ···················· | ···················· | ···················· | ···················· |
| H | 6549 hCoV-19/USA/NMDOH-2020117725/2020\|EPI_ISL_542018\|2 | ···················· | ···················· | ···················· | ···················· | ···················· | ···················· |
| H | 6550 hCoV-19/USA/NMDOH-2020117726/2020\|EPI_ISL_542019\|2 | ···················· | ···················· | ···················· | ···················· | ···················· | ···················· |
| H | 6551 hCoV-19/USA/NMDOH-2020132665/2020\|EPI_ISL_542020\|2 | ···················· | ···················· | ···················· | ···················· | ···················· | ···················· |
| H | 6552 hCoV-19/USA/NM-NMDOH-2020132668/2020\|EPI_ISL_54202 | ···················· | ···················· | ···················· | ···················· | ···················· | ···················· |
| H | 6553 hCoV-19/USA/NMDOH-2020136739/2020\|EPI_ISL_542022\|2 | ···················· | ···················· | ···················· | ···················· | ···················· | ···················· |
| H | 6554 hCoV-19/USA/NMDOH-2020153389/2020\|EPI_ISL_542023\|2 | ···················· | ···················· | ···················· | ···················· | ···················· | ···················· |
| H | 6555 hCoV-19/USA/NMDOH-2020174489/2020\|EPI_ISL_542024\|2 | ···················· | ···················· | ···················· | ···················· | ···················· | ···················· |
| H | 6556 hCoV-19/USA/NMDOH-2020174703/2020\|EPI_ISL_542025\|2 | ···················· | ···················· | ···················· | ···················· | ···················· | ···················· |
| H | 6557 hCoV-19/USA/NMDOH-2020174730/2020\|EPI_ISL_542026\|2 | ···················· | ···················· | ···················· | ···················· | ···················· | ···················· |
| H | 6558 hCoV-19/USA/NMDOH-2020174738/2020\|EPI_ISL_542027\|2 | ···················· | ···················· | ···················· | ···················· | ···················· | ···················· |
| H | 6559 hCoV-19/USA/NMDOH-2020183909/2020\|EPI_ISL_542029\|2 | ···················· | ···················· | ···················· | ···················· | ···················· | ···················· |
| H | 6560 hCoV-19/USA/NMDOH-2020185052/2020\|EPI_ISL_542030\|2 | ···················· | ···················· | ···················· | ···················· | ···················· | ···················· |
| H | 6561 hCoV-19/USA/NMDOH-2020189536/2020\|EPI_ISL_542031\|2 | ···················· | ···················· | ···················· | ···················· | ···················· | ···················· |
| H | 6562 hCoV-19/USA/NMDOH-2020189541/2020\|EPI_ISL_542032\|2 | ···················· | ···················· | ···················· | ···················· | ···················· | ···················· |
| H | 6563 hCoV-19/USA/NMDOH-2020189581/2020\|EPI_ISL_542033\|2 | ···················· | ···················· | ···················· | ···················· | ···················· | ···················· |
| H | 6564 hCoV-19/USA/NMDOH-2020189658/2020\|EPI_ISL_542035\|2 | ···················· | ···················· | ···················· | ···················· | ······T············· | ···················· |
| H | 6565 hCoV-19/USA/NMDOH-2020239972/2020\|EPI_ISL_542038\|2 | ···················· | ···················· | ···················· | ···················· | ···················· | ···················· |
| H | 6566 hCoV-19/USA/NMDOH-2020245660/2020\|EPI_ISL_542039\|2 | ···················· | ···················· | ···················· | ···················· | ···················· | ···················· |
| H | 6567 hCoV-19/USA/NMDOH-2020245669/2020\|EPI_ISL_542040\|2 | ···················· | ···················· | ···················· | ···················· | ···················· | ···················· |
| H | 6568 hCoV-19/USA/NMDOH-2020305740/2020\|EPI_ISL_542041\|2 | ···················· | ···················· | ···················· | ···················· | ···················· | ···················· |
| H | 6569 hCoV-19/USA/NMDOH-2020305741/2020\|EPI_ISL_542042\|2 | ···················· | ···················· | ···················· | ···················· | ···················· | ···················· |
| H | 6570 hCoV-19/USA/NMDOH-2020305742/2020\|EPI_ISL_542043\|2 | ···················· | ···················· | ···················· | ···················· | ···················· | ···················· |
| H | 6571 hCoV-19/USA/NMDOH-2020305743/2020\|EPI_ISL_542044\|2 | ···················· | ···················· | ···················· | ···················· | ···················· | ···················· |
| H | 6572 hCoV-19/USA/NMDOH-2020305745/2020\|EPI_ISL_542045\|2 | ···················· | ···················· | ···················· | ···················· | ···················· | ···················· |
| H | 6573 hCoV-19/USA/NMDOH-2020305746/2020\|EPI_ISL_542046\|2 | ···················· | ···················· | ···················· | ···················· | ···················· | ···················· |
| H | 6574 hCoV-19/USA/NMDOH-2020305747/2020\|EPI_ISL_542047\|2 | ···················· | ···················· | ···················· | ···················· | ···················· | ···················· |
| H | 6575 hCoV-19/USA/NMDOH-2020305749/2020\|EPI_ISL_542048\|2 | ···················· | ···················· | ···················· | ···················· | ···················· | ···················· |
| H | 6576 hCoV-19/USA/NMDOH-2020305750/2020\|EPI_ISL_542049\|2 | ···················· | ···················· | ···················· | ···················· | ···················· | ···················· |
| H | 6577 hCoV-19/USA/NMDOH-2020305755/2020\|EPI_ISL_542050\|2 | ···················· | ···················· | ···················· | ···················· | ···················· | ···················· |
| H | 6578 hCoV-19/USA/NMDOH-2020305762/2020\|EPI_ISL_542051\|2 | ···················· | ···················· | ···················· | ···················· | ···················· | ···················· |
| H | 6579 hCoV-19/USA/NMDOH-2020305763/2020\|EPI_ISL_542052\|2 | ···················· | ···················· | ···················· | ···················· | ···················· | ···················· |
| H | 6580 hCoV-19/USA/NMDOH-2020305764/2020\|EPI_ISL_542053\|2 | ···················· | ···················· | ···················· | ···················· | ···················· | ···················· |
| H | 6581 hCoV-19/USA/NMDOH-2020305771/2020\|EPI_ISL_542054\|2 | ···················· | ···················· | ···················· | ···················· | ···················· | ···················· |
| H | 6582 hCoV-19/USA/NMDOH-2020305799/2020\|EPI_ISL_542055\|2 | ···················· | ···················· | ···················· | ···················· | ···················· | ···················· |
| H | 6583 hCoV-19/USA/NMDOH-2020305802/2020\|EPI_ISL_542056\|2 | ···················· | ···················· | ···················· | ···················· | ···················· | ···················· |
| H | 6584 hCoV-19/USA/NMDOH-2020305804/2020\|EPI_ISL_542057\|2 | ···················· | ···················· | ···················· | ···················· | ···················· | ···················· |
| H | 6585 hCoV-19/USA/NMDOH-2020305805/2020\|EPI_ISL_542058\|2 | ···················· | ···················· | ···················· | ···················· | ···················· | ···················· |
| H | 6586 hCoV-19/USA/NMDOH-2020305813/2020\|EPI_ISL_542059\|2 | ···················· | ···················· | ···················· | ···················· | ···················· | ···················· |
| H | 6587 hCoV-19/USA/NMDOH-2020305819/2020\|EPI_ISL_542060\|2 | ···················· | ···················· | ···················· | ···················· | ···················· | ···················· |
| H | 6588 hCoV-19/USA/NMDOH-2020305829/2020\|EPI_ISL_542061\|2 | ···················· | ···················· | ···················· | ···················· | ···················· | ···················· |
| H | 6589 hCoV-19/USA/NMDOH-2020320036/2020\|EPI_ISL_542062\|2 | ···················· | ····A··············· | ···················· | ···················· | ···················· | ···················· |
| H | 6590 hCoV-19/USA/NMDOH-2020320930/2020\|EPI_ISL_542063\|2 | ···················· | ···················· | ···················· | ···················· | ···················· | ···················· |
| H | 6591 hCoV-19/USA/NMDOH-2020320942/2020\|EPI_ISL_542065\|2 | ···················· | ···················· | ···················· | ···················· | ···················· | ···················· |
| H | 6592 hCoV-19/USA/NMDOH-2020320943/2020\|EPI_ISL_542066\|2 | ···················· | ···················· | ···················· | ···················· | ···················· | ···················· |
| H | 6593 hCoV-19/USA/NMDOH-2020320960/2020\|EPI_ISL_542067\|2 | ···················· | ···················· | ···················· | ···················· | ···················· | ···················· |
| H | 6594 hCoV-19/USA/NMDOH-2020321601/2020\|EPI_ISL_542068\|2 | ···················· | ···················· | ···················· | ···················· | ···················· | ···················· |
| H | 6595 hCoV-19/USA/NMDOH-2020321619/2020\|EPI_ISL_542069\|2 | ···················· | ···················· | ···················· | ···················· | ···················· | ···················· |
| H | 6596 hCoV-19/USA/TX-HMH-4063/2020\|EPI_ISL_543336\|2020-0 | ···················· | ···················· | ···················· | ···················· | ···················· | ···················· |
| H | 6597 hCoV-19/USA/TX-HMH-4065/2020\|EPI_ISL_543338\|2020-0 | ···················· | ···················· | ···················· | ···················· | ···················· | ···················· |
| H | 6598 hCoV-19/USA/TX-HMH-4068/2020\|EPI_ISL_543341\|2020-0 | ···················· | ···················· | ···················· | ···················· | ···················· | ···················· |
| H | 6600 hCoV-19/USA/TX-HMH-4071/2020\|EPI_ISL_543343\|2020-0 | ···················· | ···················· | ···················· | ···················· | ···················· | ···················· |

| | | | | | | | | |
|---|---|---|---|---|---|---|---|---|
| H | 6602 hCoV-19/USA/TX-HMH-4075/2020\|EPI_ISL_543346\|2020-0 | ···················· | ···················· | ····················· | ···················· | ···················· | ···················· | |
| H | 6603 hCoV-19/USA/TX-HMH-4077/2020\|EPI_ISL_543348\|2020-0 | ···················· | ···················· | ····················· | ···················· | ···················· | ···················· | |
| H | 6606 hCoV-19/USA/TX-HMH-4083/2020\|EPI_ISL_543354\|2020-0 | ···················· | ···················· | ····················· | ···················· | ···················· | ···················· | |
| H | 6609 hCoV-19/USA/TX-HMH-4088/2020\|EPI_ISL_543358\|2020-0 | ···················· | ···················· | ····················· | ···················· | ···················· | ···················· | |
| H | 6610 hCoV-19/USA/TX-HMH-4089/2020\|EPI_ISL_543359\|2020-0 | ···················· | ···················· | ····················· | ···················· | ···················· | ···················· | |
| H | 6611 hCoV-19/USA/TX-HMH-4090/2020\|EPI_ISL_543360\|2020-0 | ···················· | ···················· | ····················· | ···················· | ···················· | ···················· | |
| H | 6612 hCoV-19/USA/TX-HMH-4091/2020\|EPI_ISL_543361\|2020-0 | ···················· | ···················· | ····················· | ···················· | ··············N····· | ···················· | |
| H | 6614 hCoV-19/USA/TX-HMH-4094/2020\|EPI_ISL_543364\|2020-0 | ···················· | ···················· | ····················· | ···················· | ···················· | ···················· | |
| H | 6615 hCoV-19/USA/TX-HMH-4097/2020\|EPI_ISL_543367\|2020-0 | ···················· | ···················· | ····················· | ···················· | ···················· | ···················· | |
| H | 6616 hCoV-19/USA/TX-HMH-4098/2020\|EPI_ISL_543368\|2020-0 | ···················· | ···················· | ····················· | ···················· | ···················· | ···················· | |
| H | 6617 hCoV-19/USA/TX-HMH-4099/2020\|EPI_ISL_543369\|2020-0 | ···················· | ···················· | ····················· | ···················· | ···················· | ···················· | |
| H | 6618 hCoV-19/USA/TX-HMH-4101/2020\|EPI_ISL_543371\|2020-0 | ···················· | ···················· | ····················· | ···················· | ···················· | ···················· | |
| H | 6619 hCoV-19/USA/TX-HMH-4102/2020\|EPI_ISL_543372\|2020-0 | ···················· | ···················· | ····················· | ···················· | ···················· | ···················· | |
| H | 6620 hCoV-19/USA/TX-HMH-4106/2020\|EPI_ISL_543374\|2020-0 | ···················· | ···················· | ····················· | ···················· | ···················· | ···················· | |
| H | 6621 hCoV-19/USA/TX-HMH-4107/2020\|EPI_ISL_543375\|2020-0 | ···················· | ···················· | ····················· | ···················· | ···················· | ···················· | |
| H | 6623 hCoV-19/USA/TX-HMH-4111/2020\|EPI_ISL_543378\|2020-0 | ···················· | ···················· | ····················· | ···················· | ···················· | N······NN··········· | 1 |
| H | 6624 hCoV-19/USA/TX-HMH-4114/2020\|EPI_ISL_543381\|2020-0 | ···················· | ···················· | ····················· | ···················· | ···················· | ···················· | |
| H | 6625 hCoV-19/USA/TX-HMH-4115/2020\|EPI_ISL_543382\|2020-0 | ···················· | ···················· | ····················· | ···················· | ···················· | ···················· | |
| H | 6626 hCoV-19/USA/TX-HMH-4119/2020\|EPI_ISL_543386\|2020-0 | ···················· | ···················· | ····················· | ···················· | ···················· | ···················· | |
| H | 6627 hCoV-19/USA/TX-HMH-4120/2020\|EPI_ISL_543387\|2020-0 | ···················· | ···················· | ····················· | ···················· | ···················· | ···················· | |
| H | 6628 hCoV-19/USA/TX-HMH-4122/2020\|EPI_ISL_543388\|2020-0 | ···················· | ···················· | ····················· | ···················· | ···················· | ···················· | |
| H | 6629 hCoV-19/USA/TX-HMH-4123/2020\|EPI_ISL_543389\|2020-0 | ···················· | ·············A······ | ····················· | ···················· | ···················· | ···················· | |
| H | 6630 hCoV-19/USA/TX-HMH-4124/2020\|EPI_ISL_543390\|2020-0 | ···················· | ···················· | ····················· | ···················· | ···················· | ···················· | |
| H | 6631 hCoV-19/USA/TX-HMH-4125/2020\|EPI_ISL_543391\|2020-0 | ···················· | ···················· | ····················· | ···················· | ···················· | ···················· | |
| H | 6632 hCoV-19/USA/TX-HMH-4128/2020\|EPI_ISL_543394\|2020-0 | ···················· | ···················· | ····················· | ···················· | ···················· | ···················· | |
| H | 6633 hCoV-19/USA/TX-HMH-4130/2020\|EPI_ISL_543395\|2020-0 | ···················· | ······A············· | ····················· | ···················· | ···················· | ···················· | |
| H | 6634 hCoV-19/USA/TX-HMH-4131/2020\|EPI_ISL_543396\|2020-0 | ···················· | ···················· | ····················· | ···················· | ···················· | ···················· | |
| H | 6635 hCoV-19/USA/TX-HMH-4132/2020\|EPI_ISL_543397\|2020-0 | ···················· | ···················· | ····················· | ···················· | ···················· | ···················· | |
| H | 6636 hCoV-19/USA/TX-HMH-4134/2020\|EPI_ISL_543398\|2020-0 | ···················· | ···················· | ····················· | ···················· | ···················· | ···················· | |
| H | 6637 hCoV-19/USA/TX-HMH-4135/2020\|EPI_ISL_543399\|2020-0 | ···················· | ···················· | ····················· | ···················· | ···················· | ···················· | |
| H | 6638 hCoV-19/USA/TX-HMH-4136/2020\|EPI_ISL_543400\|2020-0 | ···················· | ·············A······ | ····················· | ···················· | ···················· | ···················· | |
| H | 6639 hCoV-19/USA/TX-HMH-4137/2020\|EPI_ISL_543401\|2020-0 | ···················· | ···················· | ····················· | ···················· | ···················· | ···················· | |
| H | 6641 hCoV-19/USA/TX-HMH-4142/2020\|EPI_ISL_543405\|2020-0 | ···················· | ···················· | ····················· | ···················· | ···················· | ···················· | |
| H | 6642 hCoV-19/USA/TX-HMH-4167/2020\|EPI_ISL_543406\|2020-0 | ···················· | ···················· | ····················· | ···················· | ···················· | ···················· | |
| H | 6643 hCoV-19/USA/TX-HMH-4168/2020\|EPI_ISL_543407\|2020-0 | ···················· | ·············A······ | ····················· | ···················· | ···················· | ···················· | |
| H | 6645 hCoV-19/USA/TX-HMH-4170/2020\|EPI_ISL_543409\|2020-0 | ···················· | ···················· | ····················· | ···················· | ···················· | ···A················ | |
| H | 6650 hCoV-19/USA/TX-HMH-4179/2020\|EPI_ISL_543415\|2020-0 | ···················· | ···················· | ····················· | ···················· | ··············N····· | ···················· | |
| H | 6651 hCoV-19/USA/TX-HMH-4180/2020\|EPI_ISL_543416\|2020-0 | ···················· | ···················· | ····················· | ···················· | ···················· | ···················· | |
| H | 6652 hCoV-19/USA/TX-HMH-4183/2020\|EPI_ISL_543419\|2020-0 | ···················· | ···················· | ····················· | ···················· | ··············N····· | NN·····N············ | 1 |
| H | 6653 hCoV-19/USA/TX-HMH-4185/2020\|EPI_ISL_543420\|2020-0 | ···················· | ···················· | ····················· | ···················· | ···················· | ···················· | |
| H | 6654 hCoV-19/USA/TX-HMH-4187/2020\|EPI_ISL_543421\|2020-0 | ···················· | ·············A······ | ····················· | ···················· | ···················· | ···················· | |
| H | 6657 hCoV-19/England/QEUH-96BF58/2020\|EPI_ISL_531440\|20 | ···················· | ···················· | ····················· | ···················· | ···················· | ···················· | |
| H | 6658 hCoV-19/Northern Ireland/QEUH-96B978/2020\|EPI_ISL_ | ···················· | ···················· | ····················· | ···················· | ···················· | ···················· | |
| H | 6659 hCoV-19/Scotland/QEUH-96B1F8/2020\|EPI_ISL_531442\|2 | ···················· | ···················· | ····················· | ···················· | ···················· | ···················· | |
| H | 6660 hCoV-19/England/QEUH-96AE89/2020\|EPI_ISL_531443\|20 | ···················· | ·A·················· | ····················· | ···················· | ···················· | ···A················ | |
| H | 6661 hCoV-19/England/QEUH-96BCCA/2020\|EPI_ISL_531444\|20 | ···················· | ···················· | ····················· | ···················· | ···················· | ···················· | |
| H | 6662 hCoV-19/England/QEUH-96BFC1/2020\|EPI_ISL_531446\|20 | ···················· | ·A·················· | ····················· | ···················· | ···················· | ···················· | |
| H | 6663 hCoV-19/England/QEUH-96BEA6/2020\|EPI_ISL_531447\|20 | ···················· | ···················· | ····················· | ···················· | ···················· | ···················· | |
| H | 6664 hCoV-19/England/QEUH-96B231/2020\|EPI_ISL_531448\|20 | ···················· | ···················· | ····················· | ···················· | ···················· | ···················· | |
| H | 6665 hCoV-19/Northern Ireland/QEUH-96AFD1/2020\|EPI_ISL_ | ···················· | ···················· | ····················· | ···················· | ···················· | ···················· | |
| H | 6666 hCoV-19/England/QEUH-96BF0D/2020\|EPI_ISL_531450\|20 | ···················· | ···················· | ····················· | ···················· | ···················· | ···················· | |
| H | 6667 hCoV-19/England/QEUH-96B6EA/2020\|EPI_ISL_531451\|20 | ···················· | ···················· | ····················· | ···················· | ···················· | ···················· | |
| H | 6668 hCoV-19/England/QEUH-96A93D/2020\|EPI_ISL_531452\|20 | ···················· | ···················· | ····················· | ···················· | ···················· | ···················· | |
| H | 6669 hCoV-19/England/QEUH-96B987/2020\|EPI_ISL_531453\|20 | ···················· | ···················· | ····················· | ···················· | ···················· | ···················· | |
| H | 6670 hCoV-19/Northern Ireland/QEUH-96B94B/2020\|EPI_ISL_ | ···················· | ···················· | ····················· | ···················· | ···················· | ···················· | |
| H | 6671 hCoV-19/England/QEUH-96BBBE/2020\|EPI_ISL_531455\|20 | ···················· | ···················· | ····················· | ···················· | ···················· | ···················· | |
| H | 6672 hCoV-19/England/QEUH-89971E/2020\|EPI_ISL_532772\|20 | ···················· | ···················· | ····················· | ···················· | ···················· | ···················· | |
| H | 6673 hCoV-19/Northern Ireland/QEUH-8A8855/2020\|EPI_ISL_ | ···················· | ···················· | ····················· | ···················· | ···················· | ···················· | |
| H | 6674 hCoV-19/England/QEUH-89A601/2020\|EPI_ISL_532774\|20 | ···················· | ···················· | ····················· | ···················· | ···················· | ···················· | |
| H | 6675 hCoV-19/England/QEUH-883CCD/2020\|EPI_ISL_532775\|20 | ···················· | ···················· | ····················· | ···················· | ···················· | ···················· | |
| H | 6676 hCoV-19/England/QEUH-8A6B7C/2020\|EPI_ISL_532776\|20 | ···················· | ···················· | ····················· | ···················· | ···················· | ···················· | |
| H | 6677 hCoV-19/England/QEUH-881D53/2020\|EPI_ISL_532777\|20 | ···················· | ···················· | ····················· | ···················· | ···················· | ···················· | |
| H | 6678 hCoV-19/England/QEUH-88D382/2020\|EPI_ISL_532778\|20 | ···················· | ·A·················· | ····················· | ···················· | ···················· | ···················· | |
| H | 6679 hCoV-19/England/QEUH-8A03C5/2020\|EPI_ISL_532779\|20 | ···················· | ···················· | ····················· | ···················· | ···················· | ···················· | |
| H | 6680 hCoV-19/Northern Ireland/QEUH-87C82D/2020\|EPI_ISL_ | ···················· | ···················· | ····················· | ···················· | ···················· | ···················· | |
| H | 6681 hCoV-19/England/QEUH-8885FA/2020\|EPI_ISL_532783\|20 | ···················· | ···················· | ····················· | ···················· | ···················· | ···················· | |
| H | 6682 hCoV-19/Scotland/QEUH-88E415/2020\|EPI_ISL_532784\|2 | ···················· | ···················· | ····················· | ···················· | ···················· | ···················· | |
| H | 6683 hCoV-19/England/QEUH-89F411/2020\|EPI_ISL_532785\|20 | ···················· | ···················· | ····················· | ···················· | ···················· | ···················· | |
| H | 6684 hCoV-19/Scotland/QEUH-8A6523/2020\|EPI_ISL_532786\|2 | ···················· | ···················· | ····················· | ···················· | ···················· | ···················· | |
| H | 6685 hCoV-19/Northern Ireland/QEUH-7663F9/2020\|EPI_ISL_ | ···················· | ·········A·········· | ····················· | ···················· | ···················· | ···················· | |
| H | 6686 hCoV-19/England/QEUH-7689E6/2020\|EPI_ISL_533002\|20 | ···················· | ···················· | ····················· | ···················· | ···················· | ···················· | |
| H | 6687 hCoV-19/England/QEUH-75872B/2020\|EPI_ISL_533003\|20 | ···················· | ·A·················· | ····················· | ···················· | ···················· | ···················· | |
| H | 6688 hCoV-19/England/QEUH-7689AA/2020\|EPI_ISL_533004\|20 | ···················· | ···················· | ····················· | ···················· | ···················· | ···················· | |
| H | 6689 hCoV-19/England/QEUH-7606E7/2020\|EPI_ISL_533005\|20 | ···················· | ···················· | ····················· | ···················· | ···················· | ···················· | |
| H | 6690 hCoV-19/Northern Ireland/QEUH-754F54/2020\|EPI_ISL_ | ···················· | ···················· | ····················· | ···················· | ···················· | ···················· | |
| H | 6691 hCoV-19/England/QEUH-766405/2020\|EPI_ISL_533008\|20 | ···················· | ···················· | ····················· | ···················· | ···················· | ···················· | |
| H | 6692 hCoV-19/England/QEUH-7A6FEA/2020\|EPI_ISL_533009\|20 | ···················· | ···················· | ····················· | ···················· | ···················· | ···················· | |
| H | 6693 hCoV-19/England/QEUH-7A508E/2020\|EPI_ISL_533010\|20 | ···················· | ···················· | ····················· | ···················· | ···················· | ···················· | |
| H | 6694 hCoV-19/England/QEUH-795CC9/2020\|EPI_ISL_533011\|20 | ···················· | ···················· | ····················· | ···················· | ···················· | ···················· | |
| H | 6695 hCoV-19/Northern Ireland/QEUH-79CD65/2020\|EPI_ISL_ | ···················· | ···················· | ····················· | ···················· | ···················· | ···················· | |

H 6696 hCoV-19/England/QEUH-78D918/2020|EPI_ISL_533014|20 ···················· ···················· ···················· ···················· ···················· ····················
H 6697 hCoV-19/Northern Ireland/QEUH-781B1E/2020|EPI_ISL_ ···················· ···················· ···················· ···················· ···················· ····················
H 6698 hCoV-19/Scotland/QEUH-789563/2020|EPI_ISL_533016|2 ···················· ·A·················· ···················· ···················· ···················· ··A·················
H 6699 hCoV-19/England/QEUH-7923B8/2020|EPI_ISL_533018|20 ···················· ···················· ···················· ···················· ···················· ····················
H 6700 hCoV-19/England/QEUH-795FB1/2020|EPI_ISL_533019|20 ···················· ···················· ···················· ···················· ···················· ····················
H 6701 hCoV-19/England/QEUH-79ACE2/2020|EPI_ISL_533020|20 ···················· ···················· ···················· ···················· ···················· ····················
H 6702 hCoV-19/England/QEUH-7A3035/2020|EPI_ISL_533021|20 ···················· ···················· ···················· ···················· ···················· ····················
H 6703 hCoV-19/England/QEUH-78A395/2020|EPI_ISL_533022|20 ···················· ···················· ···················· ···················· ···················· ····················
H 6704 hCoV-19/England/QEUH-78634E/2020|EPI_ISL_533023|20 ···················· ···················· ···················· ···················· ···················· ····················
H 6705 hCoV-19/Northern Ireland/QEUH-77D8A5/2020|EPI_ISL_ ···················· ···················· ···················· ···················· ···················· ····················
H 6706 hCoV-19/Northern Ireland/QEUH-781A3F/2020|EPI_ISL_ ···················· ···················· ···················· ···················· ···················· ····················
H 6707 hCoV-19/Scotland/QEUH-7894C0/2020|EPI_ISL_533027|2 ···················· ·A·················· ···················· ···················· ···················· ··A·················
H 6708 hCoV-19/England/QEUH-788519/2020|EPI_ISL_533028|20 ···················· ···················· ···················· ···················· ···················· ····················
H 6709 hCoV-19/England/QEUH-788713/2020|EPI_ISL_533031|20 ···················· ···················· ···················· ···················· ···················· ····················
H 6710 hCoV-19/England/QEUH-7A7848/2020|EPI_ISL_533032|20 ···················· ···················· ···················· ···················· ···················· ····················
H 6711 hCoV-19/England/QEUH-77C41B/2020|EPI_ISL_533033|20 ···················· ···················· ···················· ···················· ···················· ····················
H 6712 hCoV-19/England/QEUH-79A20E/2020|EPI_ISL_533034|20 ···················· ···················· ···················· ···················· ···················· ····················
H 6713 hCoV-19/England/QEUH-77C976/2020|EPI_ISL_533035|20 ···················· ···················· ···················· ···················· ···················· ····················
H 6714 hCoV-19/England/QEUH-787AB1/2020|EPI_ISL_533036|20 ···················· ···················· ···················· ···················· ···················· ····················
H 6715 hCoV-19/England/QEUH-77C92B/2020|EPI_ISL_533037|20 ···················· ···················· ···················· ···················· ···················· ····················
H 6716 hCoV-19/Northern Ireland/QEUH-7A4C03/2020|EPI_ISL_ ···················· ·········A·········· ···················· ···················· ···················· ····················
H 6717 hCoV-19/England/QEUH-79202A/2020|EPI_ISL_533039|20 ···················· ···················· ···················· ···················· ···················· ····················
H 6718 hCoV-19/England/QEUH-79EEF7/2020|EPI_ISL_533040|20 ···················· ·A·················· ···················· ···················· ···················· ····················
H 6719 hCoV-19/England/QEUH-7A0979/2020|EPI_ISL_533041|20 ···················· ···················· ···················· ···················· ···················· ····················
H 6720 hCoV-19/England/QEUH-7A8FF7/2020|EPI_ISL_533042|20 ···················· ···················· ···················· ···················· ···················· ····················
H 6721 hCoV-19/England/QEUH-76CC52/2020|EPI_ISL_533043|20 ···················· ···················· ···················· ···················· ···················· ····················
H 6722 hCoV-19/England/QEUH-790709/2020|EPI_ISL_533044|20 ···················· ······A············· ···················· ···················· ···················· ····················
H 6723 hCoV-19/England/QEUH-784EA9/2020|EPI_ISL_533045|20 ···················· ···················· ···················· ···················· ···················· ····················
H 6724 hCoV-19/England/QEUH-792DE7/2020|EPI_ISL_533047|20 ···················· ···················· ···················· ···················· ···················· ····················
H 6725 hCoV-19/England/QEUH-793153/2020|EPI_ISL_533048|20 ···················· ···················· ···················· ···················· ···················· ····················
H 6726 hCoV-19/Spain/MD-H12-44/2020|EPI_ISL_534327|2020-0 ···················· ···················· ···················· ···················· ···················· ····················
H 6727 hCoV-19/Spain/MD-H12-45/2020|EPI_ISL_534328|2020-0 ···················· ·A·················· ···················· ···················· ···················· ····················
H 6728 hCoV-19/Spain/MD-H12-88/2020|EPI_ISL_534329|2020-0 ···················· ···················· ···················· ···················· ···················· ····················
H 6729 hCoV-19/Spain/MD-HLP-105-1/2020|EPI_ISL_534330|202 ···················· ···················· ···················· ···················· ···················· ····················
H 6730 hCoV-19/Spain/MD-HLP-105-2/2020|EPI_ISL_534331|202 ···················· ···················· ···················· ···················· ···················· ····················
H 6731 hCoV-19/Spain/MD-HLP-105-3/2020|EPI_ISL_534332|202 ···················· ···················· ···················· ···················· ···················· ····················
H 6732 hCoV-19/Spain/MD-HLP-105-4/2020|EPI_ISL_534333|202 ···················· ···················· ···················· ···················· ···················· ····················
H 6733 hCoV-19/Spain/MD-HRC-88/2020|EPI_ISL_534334|2020-0 ···················· ···················· ···················· ···················· ···················· ····················
H 6734 hCoV-19/Taiwan/NTU30/2020|EPI_ISL_534336|2020-08-1 ········T··········· ···················· ···················· ···················· ···················· ····················
H 6735 hCoV-19/Russia/CRIE4924360/2020|EPI_ISL_534337|202 ···················· ···················· ···················· ···················· ···················· ····················
H 6736 hCoV-19/Russia/CRIE4955821/2020|EPI_ISL_534338|202 ···················· ···················· ···················· ···················· ···················· ····················
H 6737 hCoV-19/Russia/CRIE4991898/2020|EPI_ISL_534339|202 ···················· ···················· ···················· ···················· ···················· ····················
H 6738 hCoV-19/Russia/CRIE5027243/2020|EPI_ISL_534340|202 ···················· ···················· ···················· ···················· ···················· ····················
H 6739 hCoV-19/Russia/CRIE5006606/2020|EPI_ISL_534341|202 ···················· ···················· ···················· ···················· ···················· ····················
H 6740 hCoV-19/Russia/CRIE4980432/2020|EPI_ISL_534342|202 ···················· ···················· ···················· ···················· ···················· ····················
H 6741 hCoV-19/Russia/CRIE5047362/2020|EPI_ISL_534343|202 ···················· ···················· ···················· ···················· ···················· ····················
H 6742 hCoV-19/Russia/CRIE5047007/2020|EPI_ISL_534344|202 ···················· ···················· ···················· ···················· ···················· ····················
H 6743 hCoV-19/Russia/CRIE5049919/2020|EPI_ISL_534345|202 ···················· ···················· ···················· ···················· ···················· ····················
H 6744 hCoV-19/Russia/CRIE181019/2020|EPI_ISL_534346|2020 ···················· ···················· ···················· ···················· ···················· ····················
H 6745 hCoV-19/Russia/CRIE161122/2020|EPI_ISL_534347|2020 ···················· ···················· ···················· ···················· ···················· ····················
H 6746 hCoV-19/Russia/CRIE166064/2020|EPI_ISL_534348|2020 ···················· ···················· ···················· ···················· ···················· ····················
H 6747 hCoV-19/Russia/CRIE166582/2020|EPI_ISL_534349|2020 ···················· ···················· ···················· ···················· ···················· ····················
H 6748 hCoV-19/Scotland/QEUH-964335/2020|EPI_ISL_534365|2 ···················· ···················· ···················· ···················· ···················· ····················
H 6749 hCoV-19/Scotland/QEUH-963A7D/2020|EPI_ISL_534366|2 ···················· ···················· ···················· ···················· ···················· ····················
H 6750 hCoV-19/Scotland/QEUH-963F8D/2020|EPI_ISL_534367|2 ···················· ···················· ···················· ···················· ···················· ····················
H 6751 hCoV-19/Scotland/QEUH-963FAB/2020|EPI_ISL_534368|2 ···················· ···················· ···················· ···················· ···················· ····················
H 6752 hCoV-19/Scotland/QEUH-963A6E/2020|EPI_ISL_534369|2 ···················· ···················· ···················· ···················· ···················· ····················
H 6753 hCoV-19/Scotland/QEUH-964344/2020|EPI_ISL_534371|2 ···················· ···················· ···················· ···················· ···················· ····················
H 6754 hCoV-19/Scotland/QEUH-96A126/2020|EPI_ISL_534372|2 ···················· ···················· ···················· ···················· ···················· ····················
H 6755 hCoV-19/Scotland/QEUH-964687/2020|EPI_ISL_534373|2 ···················· ···················· ···················· ···················· ···················· ····················
H 6756 hCoV-19/Scotland/QEUH-96A3A8/2020|EPI_ISL_534374|2 ···················· ·A·················· ···················· ···················· ···················· ····················
H 6757 hCoV-19/Scotland/QEUH-96512B/2020|EPI_ISL_534375|2 ···················· ···················· ···················· ···················· ···················· ····················
H 6758 hCoV-19/Scotland/QEUH-96A27E/2020|EPI_ISL_534376|2 ···················· ···················· ···················· ···················· ···················· ····················
H 6759 hCoV-19/Scotland/QEUH-963F7E/2020|EPI_ISL_534377|2 ···················· ···················· ···················· ···················· ···················· ····················
H 6760 hCoV-19/Scotland/QEUH-96A302/2020|EPI_ISL_534378|2 ···················· ···················· ···················· ···················· ···················· ····················
H 6761 hCoV-19/Scotland/QEUH-96407A/2020|EPI_ISL_534380|2 ···················· ···················· ···················· ···················· ···················· ····················
H 6762 hCoV-19/USA/AR-CDC-0866/2020|EPI_ISL_535658|2020-0 ···················· ···················· ···················· ···················· ···················· ····················
H 6763 hCoV-19/USA/AR-CDC-7905/2020|EPI_ISL_535659|2020-0 ···················· ···················· ···················· ···················· ···················· ····················
H 6764 hCoV-19/USA/AR-CDC-7920/2020|EPI_ISL_535660|2020-0 ···················· ···················· ···················· ···················· ···················· ····················
H 6765 hCoV-19/USA/AR-CDC-7940/2020|EPI_ISL_535661|2020-0 ···················· ···················· ···················· ···················· ···················· ····················
H 6766 hCoV-19/USA/CA-CDC-4684/2020|EPI_ISL_535662|2020-0 ···················· ···················· ···················· ···················· ···················· ····················
H 6767 hCoV-19/USA/CA-CDC-4685/2020|EPI_ISL_535663|2020-0 ···················· ·A·················· ···················· ···················· ···················· ····················
H 6768 hCoV-19/USA/CA-CDC-4686/2020|EPI_ISL_535664|2020-0 ···················· ·A·················· ···················· ···················· ···················· ····················
H 6769 hCoV-19/USA/CA-CDC-4687/2020|EPI_ISL_535665|2020-0 ···················· ···················· ···················· ···················· ···················· ····················
H 6770 hCoV-19/USA/CA-CDC-4688/2020|EPI_ISL_535666|2020-0 ···················· ·A·················· ···················· ···················· ···················· ····················
H 6771 hCoV-19/USA/CA-CDC-4689/2020|EPI_ISL_535667|2020-0 ···················· ···················· ···················· ···················· ···················· ····················
H 6772 hCoV-19/USA/CA-CDC-4690/2020|EPI_ISL_535668|2020-0 ···················· ···················· ···················· ···················· ···················· ····················
H 6773 hCoV-19/USA/CA-CDC-4691/2020|EPI_ISL_535669|2020-0 ···················· ·A·················· ···················· ···················· ···················· ····················
H 6774 hCoV-19/USA/CA-CDC-4693/2020|EPI_ISL_535670|2020-0 ···················· ·A·················· ···················· ···················· ···················· ····················
H 6775 hCoV-19/USA/CA-CDC-4694/2020|EPI_ISL_535671|2020-0 ···················· ···················· ···················· ···················· ···················· ····················

| | | | | | | | |
|---|---|---|---|---|---|---|---|
| H | 6776 hCoV-19/USA/CA-CDC-4695/2020\|EPI_ISL_535672\|2020-0 | .................... | .A.................. | ..................... | .................... | .................... | .................... |
| H | 6777 hCoV-19/USA/CA-CDC-4696/2020\|EPI_ISL_535673\|2020-0 | .................... | .................... | ..................... | .................... | .................... | .................... |
| H | 6778 hCoV-19/USA/CA-CDC-4697/2020\|EPI_ISL_535674\|2020-0 | .................... | .A.................. | ..................... | .................... | .................... | .................... |
| H | 6779 hCoV-19/USA/CA-CDC-4698/2020\|EPI_ISL_535675\|2020-0 | .................... | .A.................. | ..................... | .................... | .................... | .................... |
| H | 6780 hCoV-19/USA/CA-CDC-4699/2020\|EPI_ISL_535676\|2020-0 | .................... | .A.................. | ..................... | .................... | .................... | .................... |
| H | 6781 hCoV-19/USA/CA-CDC-4700/2020\|EPI_ISL_535677\|2020-0 | .................... | .A.................. | ..................... | .................... | .................... | .................... |
| H | 6782 hCoV-19/USA/CA-CDC-4701/2020\|EPI_ISL_535678\|2020-0 | .................... | .A.................. | ..................... | .................... | .................... | .................... |
| H | 6783 hCoV-19/USA/CA-CDC-4702/2020\|EPI_ISL_535679\|2020-0 | .................... | .A.................. | ..................... | .................... | .................... | .................... |
| H | 6784 hCoV-19/USA/CA-CDC-4703/2020\|EPI_ISL_535680\|2020-0 | .................... | .A.................. | ..................... | .................... | .................... | .................... |
| H | 6785 hCoV-19/USA/CA-CDC-4704/2020\|EPI_ISL_535681\|2020-0 | .................... | .A.................. | ..................... | .................... | .................... | .................... |
| H | 6786 hCoV-19/USA/CA-CDC-4705/2020\|EPI_ISL_535682\|2020-0 | .................... | .A.................. | ..................... | .................... | .................... | .................... |
| H | 6787 hCoV-19/USA/CA-CDC-4706/2020\|EPI_ISL_535683\|2020-0 | .................... | .A.................. | ..................... | .................... | .................... | .................... |
| H | 6788 hCoV-19/USA/CA-CDC-4707/2020\|EPI_ISL_535684\|2020-0 | .................... | .................... | ..................... | .................... | .................... | .................... |
| H | 6789 hCoV-19/USA/CA-CDC-4708/2020\|EPI_ISL_535685\|2020-0 | .................... | .A.................. | ..................... | .................... | .................... | .................... |
| H | 6790 hCoV-19/USA/CA-CDC-4710/2020\|EPI_ISL_535686\|2020-0 | .................... | .................... | ..................... | .................... | .................... | .................... |
| H | 6791 hCoV-19/USA/CA-CDC-4711/2020\|EPI_ISL_535687\|2020-0 | .................... | .A.................. | ..................... | .................... | .................... | .................... |
| H | 6792 hCoV-19/USA/CA-CDC-4712/2020\|EPI_ISL_535688\|2020-0 | .................... | .................... | ..................... | .................... | .................... | .................... |
| H | 6793 hCoV-19/USA/CA-CDC-4713/2020\|EPI_ISL_535689\|2020-0 | .................... | .................... | ..................... | .................... | .................... | .................... |
| H | 6794 hCoV-19/USA/CA-CDC-4714/2020\|EPI_ISL_535690\|2020-0 | .................... | .A.................. | ..................... | .................... | .................... | .................... |
| H | 6795 hCoV-19/USA/CA-CDC-4715/2020\|EPI_ISL_535691\|2020-0 | .................... | .................... | ..................... | .................... | .................... | .................... |
| H | 6796 hCoV-19/USA/CA-CDC-4716/2020\|EPI_ISL_535692\|2020-0 | .................... | .................... | ..................... | .................... | .................... | .................... |
| H | 6797 hCoV-19/USA/CA-CDC-4717/2020\|EPI_ISL_535693\|2020-0 | .................... | .................... | ..................... | .................... | .................... | .................... |
| H | 6798 hCoV-19/USA/CA-CDC-4719/2020\|EPI_ISL_535694\|2020-0 | .................... | .A.................. | ..................... | .................... | .................... | .................... |
| H | 6799 hCoV-19/USA/CA-CDC-4722/2020\|EPI_ISL_535695\|2020-0 | .................... | .A.................. | ..................... | .................... | .................... | .................... |
| H | 6800 hCoV-19/USA/CA-CDC-4724/2020\|EPI_ISL_535696\|2020-0 | .................... | .A.................. | ..................... | .................... | .................... | .................... |
| H | 6801 hCoV-19/USA/CA-CDC-4726/2020\|EPI_ISL_535697\|2020-0 | .................... | .A.................. | ..................... | .................... | .................... | .................... |
| H | 6802 hCoV-19/USA/CA-CDC-4727/2020\|EPI_ISL_535698\|2020-0 | .................... | .A.................. | ..................... | .................... | .................... | .................... |
| H | 6803 hCoV-19/USA/CA-CDC-4728/2020\|EPI_ISL_535699\|2020-0 | .................... | .A.................. | ..................... | .................... | .................... | .................... |
| H | 6804 hCoV-19/Canada/QC-LSPQ-L00239009/2020\|EPI_ISL_5362 | .................... | .................... | ..................... | .................... | .................... | .................... |
| H | 6805 hCoV-19/Canada/QC-LSPQ-L00239077/2020\|EPI_ISL_5362 | .................... | .................... | ..................... | .................... | .................... | .................... |
| H | 6806 hCoV-19/Canada/QC-LSPQ-L00239134/2020\|EPI_ISL_5362 | .................... | .................... | ..................... | .................... | .................... | .................... |
| H | 6807 hCoV-19/Canada/QC-LSPQ-L00239192/2020\|EPI_ISL_5362 | .................... | .................... | ..................... | .................... | .................... | .................... |
| H | 6808 hCoV-19/Canada/QC-LSPQ-L00239194/2020\|EPI_ISL_5362 | .................... | .................... | ..................... | .................... | .................... | .................... |
| H | 6809 hCoV-19/Canada/QC-LSPQ-L00239209/2020\|EPI_ISL_5362 | .................... | .................... | ..................... | .................... | .................... | .................... |
| H | 6810 hCoV-19/Canada/QC-LSPQ-L00239235/2020\|EPI_ISL_5362 | .................... | .................... | ..................... | .................... | .................... | .................... |
| H | 6811 hCoV-19/Canada/QC-LSPQ-L00239249/2020\|EPI_ISL_5362 | .................... | .................... | ..................... | .................... | .................... | .................... |
| H | 6812 hCoV-19/Canada/QC-LSPQ-L00239255/2020\|EPI_ISL_5362 | .................... | .................... | ..................... | .................... | .................... | .................... |
| H | 6813 hCoV-19/Canada/QC-LSPQ-L00239272/2020\|EPI_ISL_5362 | .................... | .................... | ..................... | .................... | .................... | .................... |
| H | 6814 hCoV-19/Canada/QC-LSPQ-L00239296/2020\|EPI_ISL_5362 | .................... | .................... | ..................... | .................... | .................... | .................... |
| H | 6815 hCoV-19/Canada/QC-LSPQ-L00239298/2020\|EPI_ISL_5362 | .................... | .................... | ..................... | .................... | .................... | .................... |
| H | 6816 hCoV-19/Canada/QC-LSPQ-L00239391/2020\|EPI_ISL_5362 | .................... | .................... | ..................... | .................... | .................... | .................... |
| H | 6817 hCoV-19/Canada/QC-LSPQ-L00239396/2020\|EPI_ISL_5362 | .................... | .................... | ..................... | .................... | .................... | .................... |
| H | 6818 hCoV-19/Canada/QC-LSPQ-L00239397/2020\|EPI_ISL_5362 | .................... | .................... | ..................... | .................... | .................... | .................... |
| H | 6819 hCoV-19/Canada/QC-LSPQ-L00239402/2020\|EPI_ISL_5362 | .................... | .................... | ..................... | .................... | .................... | .................... |
| H | 6820 hCoV-19/Canada/QC-LSPQ-L00239407/2020\|EPI_ISL_5362 | .................... | .................... | ..................... | .................... | .................... | .................... |
| H | 6821 hCoV-19/Canada/QC-LSPQ-L00239412/2020\|EPI_ISL_5362 | .................... | .................... | ..................... | .................... | .................... | .................... |
| H | 6822 hCoV-19/Canada/QC-LSPQ-L00239413/2020\|EPI_ISL_5362 | .................... | .................... | ..................... | .................... | .................... | .................... |
| H | 6823 hCoV-19/Canada/QC-LSPQ-L00239865/2020\|EPI_ISL_5362 | .................... | .................... | ..................... | .................... | .................... | .................... |
| H | 6824 hCoV-19/Canada/QC-LSPQ-L00239991/2020\|EPI_ISL_5362 | .................... | .................... | ..................... | .................... | .................... | .................... |
| H | 6825 hCoV-19/Canada/QC-LSPQ-L00240197/2020\|EPI_ISL_5362 | .................... | .................... | ..................... | .................... | .................... | .................... |
| H | 6826 hCoV-19/Canada/QC-LSPQ-L00240292/2020\|EPI_ISL_5362 | .................... | .................... | ..................... | .................... | .................... | .................... |
| H | 6827 hCoV-19/Canada/QC-LSPQ-L00240404/2020\|EPI_ISL_5362 | .................... | .................... | ..................... | .................... | .................... | .................... |
| H | 6828 hCoV-19/Canada/QC-LSPQ-L00240414/2020\|EPI_ISL_5362 | .................... | .................... | ..................... | .................... | .................... | .................... |
| H | 6829 hCoV-19/Canada/QC-LSPQ-L00240479/2020\|EPI_ISL_5362 | .................... | .................... | ..................... | .................... | .................... | .................... |
| H | 6830 hCoV-19/Canada/QC-LSPQ-L00240536/2020\|EPI_ISL_5362 | .................... | .................... | ..................... | .................... | .................... | .................... |
| H | 6831 hCoV-19/Canada/QC-LSPQ-L00240552/2020\|EPI_ISL_5362 | .................... | .................... | ..................... | .................... | .................... | .................... |
| H | 6832 hCoV-19/Canada/QC-LSPQ-L00241242/2020\|EPI_ISL_5362 | .................... | .................... | ..................... | .................... | .................... | .................... |
| H | 6833 hCoV-19/Canada/QC-LSPQ-L00241433/2020\|EPI_ISL_5362 | .................... | .................... | ..................... | .................... | .................... | .................... |
| H | 6834 hCoV-19/Canada/QC-LSPQ-L00241435/2020\|EPI_ISL_5362 | .................... | .................... | ..................... | .................... | .................... | .................... |
| H | 6835 hCoV-19/Canada/QC-LSPQ-L00241439/2020\|EPI_ISL_5362 | .................... | .................... | ..................... | .................... | .................... | .................... |
| H | 6836 hCoV-19/Canada/QC-LSPQ-L00241444/2020\|EPI_ISL_5362 | .................... | .................... | ..................... | .................... | .................... | .................... |
| H | 6837 hCoV-19/Canada/QC-LSPQ-L00241514/2020\|EPI_ISL_5362 | .................... | .................... | ..................... | .................... | .................... | .................... |
| H | 6838 hCoV-19/Canada/QC-LSPQ-L00241602/2020\|EPI_ISL_5362 | .................... | .................... | ..................... | .................... | .................... | .................... |
| H | 6839 hCoV-19/Canada/QC-LSPQ-L00241610/2020\|EPI_ISL_5362 | .................... | .................... | ..................... | .................... | .................... | .................... |
| H | 6840 hCoV-19/Canada/QC-LSPQ-L00241648/2020\|EPI_ISL_5362 | .................... | .................... | ..................... | .................... | .................... | .................... |
| H | 6841 hCoV-19/Canada/QC-LSPQ-L00241670/2020\|EPI_ISL_5362 | .................... | .................... | ..................... | .................... | .................... | .................... |
| H | 6842 hCoV-19/Canada/QC-LSPQ-L00241753/2020\|EPI_ISL_5362 | .................... | .................... | ..................... | .................... | .................... | .................... |
| H | 6843 hCoV-19/Canada/QC-LSPQ-L00241782/2020\|EPI_ISL_5362 | .................... | .................... | ..................... | .................... | .................... | .................... |
| H | 6844 hCoV-19/Canada/QC-LSPQ-L00241862/2020\|EPI_ISL_5362 | .................... | .................... | ..................... | .................... | .................... | .................... |
| H | 6845 hCoV-19/Canada/QC-LSPQ-L00241900/2020\|EPI_ISL_5362 | .................... | .................... | ..................... | .................... | .................... | .................... |
| H | 6846 hCoV-19/Canada/QC-LSPQ-L00241901/2020\|EPI_ISL_5362 | .................... | .................... | ..................... | .................... | .................... | .................... |
| H | 6847 hCoV-19/Canada/QC-LSPQ-L00241907/2020\|EPI_ISL_5362 | .................... | .................... | ..................... | .................... | .................... | .................... |
| H | 6848 hCoV-19/Canada/QC-LSPQ-L00241909/2020\|EPI_ISL_5362 | .................... | .................... | ..................... | .................... | .................... | .................... |
| H | 6849 hCoV-19/Canada/QC-LSPQ-L00241924/2020\|EPI_ISL_5362 | .................... | .................... | ..................... | .................... | .................... | .................... |
| H | 6850 hCoV-19/Canada/QC-LSPQ-L00241945/2020\|EPI_ISL_5362 | .................... | .................... | ..................... | .................... | .................... | .................... |
| H | 6851 hCoV-19/Canada/QC-LSPQ-L00241948/2020\|EPI_ISL_5362 | .................... | .................... | ..................... | .................... | .................... | .................... |
| H | 6852 hCoV-19/Canada/QC-LSPQ-L00241959/2020\|EPI_ISL_5362 | .................... | .................... | ..................... | .................... | .................... | .................... |
| H | 6853 hCoV-19/England/QEUH-961BA9/2020\|EPI_ISL_536989\|20 | .................... | .................... | ..................... | .................... | .................... | .................... |
| H | 6854 hCoV-19/England/QEUH-95E0A0/2020\|EPI_ISL_536990\|20 | .................... | .................... | ..................... | .................... | .................... | .................... |
| H | 6855 hCoV-19/England/QEUH-95E40E/2020\|EPI_ISL_536991\|20 | .................... | .................... | ..................... | .................... | .................... | .................... |

| | | | | | | | | | |
|---|---|---|---|---|---|---|---|---|---|
| H | 6856 | hCoV-19/Northern Ireland/QEUH-961A51/2020\|EPI_ISL_ | ···················· | ···················· | ···················· | ···················· | ···················· | ···················· | |
| H | 6857 | hCoV-19/Northern Ireland/QEUH-961BC7/2020\|EPI_ISL_ | ···················· | ···················· | ···················· | ···················· | ···················· | ···················· | |
| H | 6858 | hCoV-19/England/QEUH-95DD8C/2020\|EPI_ISL_536996\|20 | ···················· | ···················· | ···················· | ···················· | ···················· | ···················· | |
| H | 6859 | hCoV-19/England/QEUH-96171E/2020\|EPI_ISL_536997\|20 | ···················· | ···················· | ···················· | ···················· | ···················· | ···················· | |
| H | 6860 | hCoV-19/England/QEUH-961848/2020\|EPI_ISL_536998\|20 | ···················· | ···················· | ···················· | ···················· | ···················· | ···················· | |
| H | 6861 | hCoV-19/England/QEUH-96174B/2020\|EPI_ISL_536999\|20 | ···················· | ···················· | ···················· | ···················· | ···················· | ···················· | |
| H | 6862 | hCoV-19/USA/CA-UCLA-28/2020\|EPI_ISL_537560\|2020-05 | ···················· | ·A·················· | ···················· | ···················· | ···················· | ···················· | |
| H | 6863 | hCoV-19/USA/CA-UCLA-29/2020\|EPI_ISL_537561\|2020-05 | ···················· | ···················· | ···················· | ···················· | ···················· | ···················· | |
| H | 6864 | hCoV-19/USA/CA-UCLA-3/2020\|EPI_ISL_537562\|2020-04- | ···················· | ···················· | ···················· | ···················· | ···················· | ···················· | |
| H | 6865 | hCoV-19/USA/CA-UCLA-30/2020\|EPI_ISL_537563\|2020-05 | ···················· | ···················· | ···················· | ···················· | ···················· | ···················· | |
| H | 6866 | hCoV-19/USA/CA-UCLA-31/2020\|EPI_ISL_537564\|2020-06 | ···················· | ·A·················· | ···················· | ···················· | ···················· | ···················· | |
| H | 6867 | hCoV-19/USA/CA-UCLA-32/2020\|EPI_ISL_537565\|2020-06 | ···················· | ·····A·············· | ···················· | ···················· | ···················· | ···················· | |
| H | 6868 | hCoV-19/USA/CA-UCLA-33/2020\|EPI_ISL_537566\|2020-06 | ···················· | ···················· | ···················· | ···················· | ···················· | ···················· | |
| H | 6869 | hCoV-19/USA/CA-UCLA-34/2020\|EPI_ISL_537567\|2020-06 | ···················· | ···················· | ···················· | ···················· | ···················· | ···················· | |
| H | 6870 | hCoV-19/USA/CA-UCLA-36/2020\|EPI_ISL_537568\|2020-03 | ···················· | ···················· | ···················· | ···················· | ···················· | ···················· | |
| H | 6871 | hCoV-19/USA/CA-UCLA-37/2020\|EPI_ISL_537569\|2020-03 | ···················· | ···················· | ···················· | ···················· | ···················· | ···················· | |
| H | 6872 | hCoV-19/USA/CA-UCLA-4/2020\|EPI_ISL_537570\|2020-04- | ···················· | ···················· | ···················· | ···················· | ···················· | ···················· | |
| H | 6873 | hCoV-19/USA/CA-UCLA-40/2020\|EPI_ISL_537571\|2020-03 | ···················· | ···················· | ···················· | ···················· | ···················· | ···················· | |
| H | 6874 | hCoV-19/USA/CA-UCLA-44/2020\|EPI_ISL_537573\|2020-03 | ···················· | ···················· | ···················· | ···················· | ···················· | ···················· | |
| H | 6875 | hCoV-19/USA/CA-UCLA-48/2020\|EPI_ISL_537577\|2020-03 | ···················· | ···················· | ···················· | ···················· | ···················· | ···················· | |
| H | 6876 | hCoV-19/USA/CA-UCLA-49/2020\|EPI_ISL_537578\|2020-03 | ···················· | ···················· | ···················· | ···················· | ···················· | ···················· | |
| H | 6877 | hCoV-19/USA/CA-UCLA-5/2020\|EPI_ISL_537579\|2020-04- | ···················· | ···················· | ···················· | ···················· | ···················· | ···················· | |
| H | 6878 | hCoV-19/USA/CA-UCLA-56/2020\|EPI_ISL_537582\|2020-03 | ···················· | ···················· | ···················· | ···················· | ···················· | ···················· | |
| H | 6879 | hCoV-19/USA/CA-UCLA-57/2020\|EPI_ISL_537583\|2020-03 | ···················· | ···················· | ···················· | ···················· | ···················· | ···················· | |
| H | 6880 | hCoV-19/USA/CA-UCLA-59/2020\|EPI_ISL_537584\|2020-03 | ···················· | ···················· | ···················· | ···················· | ···················· | ···················· | |
| H | 6881 | hCoV-19/USA/CA-UCLA-6/2020\|EPI_ISL_537585\|2020-04- | ···················· | ···················· | ···················· | ···················· | ···················· | ···················· | |
| H | 6882 | hCoV-19/USA/CA-UCLA-60/2020\|EPI_ISL_537586\|2020-03 | ···················· | ···················· | ···················· | ···················· | ···················· | ···················· | |
| H | 6883 | hCoV-19/USA/CA-UCLA-62/2020\|EPI_ISL_537587\|2020-03 | ···················· | ···················· | ···················· | ···················· | ···················· | ···················· | |
| H | 6884 | hCoV-19/USA/CA-UCLA-65/2020\|EPI_ISL_537590\|2020-03 | ···················· | ···················· | ···················· | ···················· | ···················· | ···················· | |
| H | 6885 | hCoV-19/USA/CA-UCLA-66/2020\|EPI_ISL_537591\|2020-03 | ···················· | ···················· | ···················· | ···················· | ···················· | ···················· | |
| H | 6886 | hCoV-19/USA/CA-UCLA-67/2020\|EPI_ISL_537592\|2020-04 | ···················· | ···················· | ···················· | ···················· | ···················· | ···················· | |
| H | 6887 | hCoV-19/USA/CA-UCLA-68/2020\|EPI_ISL_537593\|2020-04 | ···················· | ···················· | ···················· | ···················· | ···················· | ···················· | |
| H | 6888 | hCoV-19/USA/CA-UCLA-7/2020\|EPI_ISL_537594\|2020-04- | ···················· | ···················· | ···················· | ···················· | ···················· | ···················· | |
| H | 6889 | hCoV-19/USA/CA-UCLA-70/2020\|EPI_ISL_537595\|2020-03 | ···················· | ···················· | ···················· | ···················· | ···················· | ···················· | |
| H | 6890 | hCoV-19/USA/CA-UCLA-71/2020\|EPI_ISL_537596\|2020-03 | ···················· | ···················· | ···················· | ···················· | ···················· | ···················· | |
| H | 6891 | hCoV-19/USA/CA-UCLA-72/2020\|EPI_ISL_537597\|2020-03 | ···················· | ···················· | ···················· | ···················· | ···················· | ···················· | |
| H | 6892 | hCoV-19/USA/CA-UCLA-77/2020\|EPI_ISL_537598\|2020-03 | ···················· | ···················· | ···················· | ···················· | ···················· | ···················· | |
| H | 6893 | hCoV-19/USA/CA-UCLA-78/2020\|EPI_ISL_537599\|2020-03 | ···················· | ···················· | ···················· | ···················· | ···················· | ···················· | |
| H | 6894 | hCoV-19/England/LEED-2AA2F1/2020\|EPI_ISL_538890\|20 | ···················· | ···················· | ···················· | ···················· | ···················· | ···················· | |
| H | 6895 | hCoV-19/England/LEED-2A99B1/2020\|EPI_ISL_538891\|20 | ···················· | ···················· | ···················· | ···················· | ···················· | ···················· | |
| H | 6896 | hCoV-19/England/LEED-2AA4A0/2020\|EPI_ISL_538892\|20 | ···················· | ·A·················· | ···················· | ···················· | ···················· | ···················· | |
| H | 6897 | hCoV-19/England/LEED-2AA5CB/2020\|EPI_ISL_538893\|20 | ···················· | ···················· | ···················· | ···················· | ···················· | ···················· | |
| H | 6898 | hCoV-19/England/LEED-2AA631/2020\|EPI_ISL_538894\|20 | ···················· | ···················· | ···················· | ···················· | ···················· | ···················· | |
| H | 6899 | hCoV-19/England/LEED-2AA1D6/2020\|EPI_ISL_538895\|20 | ···················· | ···A················ | ···················· | ···················· | ···················· | ···················· | |
| H | 6900 | hCoV-19/England/LEED-2AA0AC/2020\|EPI_ISL_538896\|20 | ···················· | ···················· | ···················· | ···················· | ···················· | ···················· | |
| H | 6901 | hCoV-19/England/LEED-2AA376/2020\|EPI_ISL_538897\|20 | ···················· | ···················· | ···················· | ···················· | ···················· | ···················· | |
| H | 6902 | hCoV-19/Italy/LOM-UniMI-L102/2020\|EPI_ISL_542100\|2 | ···················· | ···················· | ···················· | ···················· | ···················· | ···················· | |
| H | 6903 | hCoV-19/Italy/LOM-UniMI-L103/2020\|EPI_ISL_542101\|2 | ···················· | ···················· | ···················· | ···················· | ···················· | ···················· | |
| H | 6904 | hCoV-19/Italy/LOM-UniMI-L104/2020\|EPI_ISL_542102\|2 | ···················· | ···················· | ···················· | ···················· | ···················· | ···················· | |
| H | 6905 | hCoV-19/Italy/LOM-UniMI-L105/2020\|EPI_ISL_542103\|2 | ···················· | ···················· | ···················· | ···················· | ···················· | ···················· | |
| H | 6907 | hCoV-19/USA/TX-HMH-4194/2020\|EPI_ISL_543427\|2020-0 | ···················· | ···················· | ···················· | ···················· | ···················· | ···················· | |
| H | 6908 | hCoV-19/USA/TX-HMH-4198/2020\|EPI_ISL_543429\|2020-0 | ···················· | ···················· | ···················· | ···················· | ···················· | ···N···NN··········· | 1 |
| H | 6909 | hCoV-19/USA/TX-HMH-4199/2020\|EPI_ISL_543430\|2020-0 | ···················· | ···················· | ···················· | ···················· | ···················· | ···················· | |
| H | 6910 | hCoV-19/USA/TX-HMH-4200/2020\|EPI_ISL_543431\|2020-0 | ···················· | ·A·················· | ···················· | ···················· | ···················· | ···················· | |
| H | 6911 | hCoV-19/USA/TX-HMH-4202/2020\|EPI_ISL_543433\|2020-0 | ···················· | ···················· | ···················· | ···················· | ···················· | ···················· | |
| H | 6912 | hCoV-19/USA/TX-HMH-4203/2020\|EPI_ISL_543434\|2020-0 | ···················· | ···················· | ···················· | ···················· | ···················· | ···················· | |
| H | 6913 | hCoV-19/Northern Ireland/QEUH-969D8D/2020\|EPI_ISL_ | ···················· | ···················· | ···················· | ···················· | ···················· | ···················· | |
| H | 6914 | hCoV-19/England/QEUH-96C69E/2020\|EPI_ISL_530885\|20 | ···················· | ···················· | ···················· | ···················· | ···················· | ···················· | |
| H | 6915 | hCoV-19/England/QEUH-96C555/2020\|EPI_ISL_530886\|20 | ···················· | ···················· | ···················· | ···················· | ···················· | ···················· | |
| H | 6916 | hCoV-19/England/QEUH-969F0F/2020\|EPI_ISL_530887\|20 | ···················· | ···················· | ···················· | ···················· | ···················· | ···················· | |
| H | 6917 | hCoV-19/Northern Ireland/QEUH-96C5DD/2020\|EPI_ISL_ | ···················· | ···················· | ···················· | ···················· | ···················· | ···················· | |
| H | 6918 | hCoV-19/Northern Ireland/QEUH-96C31F/2020\|EPI_ISL_ | ···················· | ···················· | ···················· | ···················· | ···················· | ···················· | |
| H | 6919 | hCoV-19/England/QEUH-969FD2/2020\|EPI_ISL_530891\|20 | ···················· | ···················· | ···················· | ···················· | ···················· | ···················· | |
| H | 6920 | hCoV-19/England/QEUH-969CEA/2020\|EPI_ISL_530892\|20 | ···················· | ···················· | ···················· | ···················· | ···················· | ···················· | |
| H | 6921 | hCoV-19/Northern Ireland/QEUH-969DD8/2020\|EPI_ISL_ | ···················· | ···················· | ···················· | ···················· | ···················· | ···················· | |
| H | 6922 | hCoV-19/Northern Ireland/QEUH-969D14/2020\|EPI_ISL_ | ···················· | ·A·················· | ···················· | ···················· | ···················· | ···················· | |
| H | 6923 | hCoV-19/England/QEUH-969CAE/2020\|EPI_ISL_530895\|20 | ···················· | ···················· | ···················· | ···················· | ···················· | ···················· | |
| H | 6924 | hCoV-19/England/QEUH-76ABFD/2020\|EPI_ISL_533050\|20 | ···················· | ···················· | ···················· | ···················· | ···················· | ···················· | |
| H | 6925 | hCoV-19/England/QEUH-7A7E55/2020\|EPI_ISL_533051\|20 | ···················· | ···················· | ···················· | ···················· | ···················· | ···················· | |
| H | 6926 | hCoV-19/Northern Ireland/QEUH-77A78D/2020\|EPI_ISL_ | ···················· | ···················· | ···················· | ···················· | ···················· | ···················· | |
| H | 6927 | hCoV-19/England/QEUH-79F351/2020\|EPI_ISL_533053\|20 | ···················· | ···················· | ···················· | ···················· | ···················· | ···················· | |
| H | 6928 | hCoV-19/England/QEUH-7A2EE1/2020\|EPI_ISL_533056\|20 | ···················· | ···················· | ···················· | ···················· | ···················· | ···················· | |
| H | 6929 | hCoV-19/England/QEUH-788E69/2020\|EPI_ISL_533057\|20 | ···················· | ···················· | ···················· | ···················· | ···················· | ···················· | |
| H | 6930 | hCoV-19/England/QEUH-7987C5/2020\|EPI_ISL_533058\|20 | ···················· | ···················· | ···················· | ···················· | ···················· | ···················· | |
| H | 6931 | hCoV-19/England/QEUH-7A4BBB/2020\|EPI_ISL_533060\|20 | ···················· | ···················· | ···················· | ···················· | ···················· | ···················· | |
| H | 6932 | hCoV-19/England/QEUH-797C03/2020\|EPI_ISL_533061\|20 | ···················· | ···················· | ···················· | ···················· | ···················· | ···················· | |
| H | 6933 | hCoV-19/England/QEUH-78498A/2020\|EPI_ISL_533062\|20 | ···················· | ···················· | ···················· | ···················· | ···················· | ···················· | |
| H | 6934 | hCoV-19/England/QEUH-788986/2020\|EPI_ISL_533063\|20 | ···················· | ···················· | ···················· | ···················· | ···················· | ···················· | |
| H | 6935 | hCoV-19/England/QEUH-789F0B/2020\|EPI_ISL_533064\|20 | ···················· | ···················· | ···················· | ···················· | ···················· | ···················· | |
| H | 6936 | hCoV-19/Northern Ireland/QEUH-79CA40/2020\|EPI_ISL_ | ···················· | ···················· | ···················· | ···················· | ···················· | ···················· | |

| | | | | | | | | |
|---|---|---|---|---|---|---|---|---|
| H | 6937 | hCoV-19/Northern Ireland/QEUH-77BD4E/2020\|EPI_ISL_ | ···················· | ···················· | ···················· | ···················· | ···················· | ···················· |
| H | 6938 | hCoV-19/England/QEUH-788889/2020\|EPI_ISL_533068\|20 | ···················· | ·A·················· | ···················· | ···················· | ···················· | ···················· |
| H | 6939 | hCoV-19/England/QEUH-77D429/2020\|EPI_ISL_533069\|20 | ···················· | ·········A·········· | ···················· | ···················· | ···················· | ···················· |
| H | 6940 | hCoV-19/Northern Ireland/QEUH-79A338/2020\|EPI_ISL_ | ···················· | ···················· | ···················· | ···················· | ···················· | ···················· |
| H | 6941 | hCoV-19/England/QEUH-7A08D6/2020\|EPI_ISL_533071\|20 | ···················· | ···················· | ···················· | ···················· | ···················· | ···················· |
| H | 6942 | hCoV-19/Scotland/QEUH-7A4553/2020\|EPI_ISL_533072\|2 | ···················· | ···················· | ···················· | ···················· | ···················· | ···················· |
| H | 6943 | hCoV-19/England/QEUH-795FDF/2020\|EPI_ISL_533073\|20 | ···················· | ···················· | ···················· | ···················· | ···················· | ···················· |
| H | 6944 | hCoV-19/Northern Ireland/QEUH-77D7B7/2020\|EPI_ISL_ | ···················· | ···················· | ···················· | ···················· | ···················· | ···················· |
| H | 6945 | hCoV-19/England/QEUH-793663/2020\|EPI_ISL_533075\|20 | ···················· | ···················· | ···················· | ···················· | ···················· | ···················· |
| H | 6946 | hCoV-19/England/QEUH-78CC4D/2020\|EPI_ISL_533076\|20 | ···················· | ···················· | ···················· | ···················· | ···················· | ···················· |
| H | 6947 | hCoV-19/Northern Ireland/QEUH-79CC95/2020\|EPI_ISL_ | ···················· | ···················· | ···················· | ···················· | ···················· | ···················· |
| H | 6948 | hCoV-19/England/QEUH-76E0BE/2020\|EPI_ISL_533078\|20 | ···················· | ···················· | ···················· | ···················· | ···················· | ···················· |
| H | 6949 | hCoV-19/Brazil/SP-309/2020\|EPI_ISL_534311\|2020-04- | ···················· | ···················· | ···················· | ···················· | ···················· | ···················· |
| H | 6950 | hCoV-19/Brazil/SP-315/2020\|EPI_ISL_534312\|2020-05- | ···················· | ···················· | ···················· | ···················· | ···················· | ···················· |
| H | 6951 | hCoV-19/Brazil/SP-316/2020\|EPI_ISL_534313\|2020-04- | ···················· | ···················· | ···················· | ···················· | ···················· | ···················· |
| H | 6952 | hCoV-19/Brazil/SP-317/2020\|EPI_ISL_534314\|2020-05- | ···················· | ···················· | ···················· | ···················· | ···················· | ···················· |
| H | 6953 | hCoV-19/Brazil/SP-318/2020\|EPI_ISL_534315\|2020-05- | ···················· | ···················· | ···················· | ···················· | ···················· | ···················· |
| H | 6954 | hCoV-19/Brazil/SP-321/2020\|EPI_ISL_534316\|2020-05- | ···················· | ···················· | ···················· | ···················· | ···················· | ···················· |
| H | 6955 | hCoV-19/Brazil/SP-324/2020\|EPI_ISL_534317\|2020-05- | ···················· | ···················· | ···················· | ···················· | ···················· | ···················· |
| H | 6956 | hCoV-19/Brazil/SP-325/2020\|EPI_ISL_534318\|2020-05- | ···················· | ···················· | ···················· | ···················· | ···················· | ···················· |
| H | 6957 | hCoV-19/Brazil/SP-255/2020\|EPI_ISL_534319\|2020-05- | ···················· | ···················· | ···················· | ···················· | ···················· | ···················· |
| H | 6958 | hCoV-19/Brazil/SP-256/2020\|EPI_ISL_534320\|2020-05- | ···················· | ···················· | ···················· | ···················· | ···················· | ···················· |
| H | 6959 | hCoV-19/Brazil/SP-329/2020\|EPI_ISL_534321\|2020-05- | ···················· | ···················· | ···················· | ···················· | ···················· | ···················· |
| H | 6960 | hCoV-19/Brazil/SP-302/2020\|EPI_ISL_534322\|2020-04- | ···················· | ···················· | ···················· | ···················· | ···················· | ···················· |
| H | 6961 | hCoV-19/Brazil/SP-304/2020\|EPI_ISL_534323\|2020-04- | ···················· | ···················· | ···················· | ···················· | ···················· | ···················· |
| H | 6962 | hCoV-19/Brazil/SP-305/2020\|EPI_ISL_534324\|2020-04- | ···················· | ···················· | ···················· | ···················· | ···················· | ···················· |
| H | 6963 | hCoV-19/Brazil/SP-306/2020\|EPI_ISL_534325\|2020-04- | ···················· | ···················· | ···················· | ···················· | ···················· | ···················· |
| H | 6964 | hCoV-19/Brazil/SP-340/2020\|EPI_ISL_534326\|2020-05- | ···················· | ···················· | ···················· | ···················· | ···················· | ···················· |
| H | 6965 | hCoV-19/Scotland/QEUH-96A092/2020\|EPI_ISL_534382\|2 | ···················· | ···················· | ···················· | ···················· | ···················· | ···················· |
| H | 6966 | hCoV-19/Scotland/QEUH-964757/2020\|EPI_ISL_534383\|2 | ···················· | ···················· | ···················· | ···················· | ···················· | ···················· |
| H | 6967 | hCoV-19/Scotland/QEUH-963A8C/2020\|EPI_ISL_534384\|2 | ···················· | ···················· | ···················· | ···················· | ···················· | ···················· |
| H | 6968 | hCoV-19/Scotland/QEUH-964195/2020\|EPI_ISL_534385\|2 | ···················· | ···················· | ···················· | ···················· | ···················· | ···················· |
| H | 6969 | hCoV-19/Scotland/QEUH-96A135/2020\|EPI_ISL_534386\|2 | ···················· | ···················· | ···················· | ···················· | ···················· | ···················· |
| H | 6970 | hCoV-19/Scotland/QEUH-964739/2020\|EPI_ISL_534387\|2 | ···················· | ···················· | ···················· | ···················· | ···················· | ···················· |
| H | 6971 | hCoV-19/Scotland/QEUH-9644D8/2020\|EPI_ISL_534389\|2 | ···················· | ···················· | ···················· | ···················· | ···················· | ···················· |
| H | 6972 | hCoV-19/Scotland/QEUH-969FF0/2020\|EPI_ISL_534390\|2 | ···················· | ···················· | ···················· | ···················· | ···················· | ···················· |
| H | 6973 | hCoV-19/Scotland/QEUH-963F50/2020\|EPI_ISL_534391\|2 | ···················· | ···················· | ···················· | ···················· | ···················· | ···················· |
| H | 6974 | hCoV-19/Scotland/QEUH-9640C5/2020\|EPI_ISL_534392\|2 | ···················· | ···················· | ···················· | ···················· | ···················· | ···················· |
| H | 6975 | hCoV-19/Scotland/QEUH-9644C9/2020\|EPI_ISL_534393\|2 | ···················· | ···················· | ···················· | ···················· | ···················· | ···················· |
| H | 6976 | hCoV-19/Scotland/QEUH-964748/2020\|EPI_ISL_534394\|2 | ···················· | ···················· | ···················· | ···················· | ···················· | ···················· |
| H | 6977 | hCoV-19/Scotland/QEUH-9644E7/2020\|EPI_ISL_534396\|2 | ···················· | ···················· | ···················· | ···················· | ···················· | ···················· |
| H | 6978 | hCoV-19/Scotland/QEUH-9647A2/2020\|EPI_ISL_534397\|2 | ···················· | ···················· | ···················· | ···················· | ···················· | ···················· |
| H | 6979 | hCoV-19/Scotland/QEUH-964308/2020\|EPI_ISL_534398\|2 | ···················· | ···················· | ···················· | ···················· | ···················· | ···················· |
| H | 6980 | hCoV-19/Scotland/QEUH-96A28D/2020\|EPI_ISL_534399\|2 | ···················· | ···················· | ···················· | ···················· | ···················· | ···················· |
| H | 6981 | hCoV-19/Switzerland/BL-ETHZ-250077/2020\|EPI_ISL_53 | ···················· | ···················· | ···················· | ···················· | ···················· | ···················· |
| H | 6982 | hCoV-19/Switzerland/GR-ETHZ-250080/2020\|EPI_ISL_53 | ···················· | ···················· | ···················· | ···················· | ···················· | ···················· |
| H | 6983 | hCoV-19/Switzerland/BE-ETHZ-250085/2020\|EPI_ISL_53 | ···················· | ···················· | ···················· | ···················· | ···················· | ···················· |
| H | 6984 | hCoV-19/Switzerland/BL-ETHZ-250087/2020\|EPI_ISL_53 | ···················· | ···················· | ···················· | ···················· | ···················· | ···················· |
| H | 6985 | hCoV-19/Switzerland/BL-ETHZ-250088/2020\|EPI_ISL_53 | ···················· | ·A·················· | ···················· | ···················· | ···················· | ···················· |
| H | 6986 | hCoV-19/Switzerland/BL-ETHZ-250090/2020\|EPI_ISL_53 | ···················· | ···················· | ···················· | ···················· | ···················· | ···················· |
| H | 6987 | hCoV-19/Switzerland/BS-ETHZ-250092/2020\|EPI_ISL_53 | ···················· | ···················· | ···················· | ···················· | ···················· | ···················· |
| H | 6988 | hCoV-19/Switzerland/BE-ETHZ-250096/2020\|EPI_ISL_53 | ···················· | ···················· | ···················· | ···················· | ···················· | ···················· |
| H | 6989 | hCoV-19/Switzerland/BE-ETHZ-250097/2020\|EPI_ISL_53 | ···················· | ···················· | ···················· | ···················· | ···················· | ···················· |
| H | 6990 | hCoV-19/Switzerland/BE-ETHZ-250098/2020\|EPI_ISL_53 | ···················· | ···················· | ···················· | ···················· | ···················· | ···················· |
| H | 6991 | hCoV-19/Switzerland/ZH-ETHZ-250099/2020\|EPI_ISL_53 | ···················· | ···················· | ···················· | ···················· | ···················· | ···················· |
| H | 6992 | hCoV-19/Switzerland/VS-ETHZ-250103/2020\|EPI_ISL_53 | ···················· | ···················· | ···················· | ···················· | ···················· | ···················· |
| H | 6993 | hCoV-19/Switzerland/BL-ETHZ-250106/2020\|EPI_ISL_53 | ···················· | ···················· | ···················· | ···················· | ···················· | ···················· |
| H | 6994 | hCoV-19/Switzerland/ZH-ETHZ-250108/2020\|EPI_ISL_53 | ···················· | ···················· | ···················· | ···················· | ···················· | ···················· |
| H | 6995 | hCoV-19/Switzerland/AG-ETHZ-250115/2020\|EPI_ISL_53 | ···················· | ·A·················· | ···················· | ···················· | ···················· | ···················· |
| H | 6996 | hCoV-19/Switzerland/BE-ETHZ-250116/2020\|EPI_ISL_53 | ···················· | ···················· | ···················· | ···················· | ······K············· | ···················· |
| H | 6997 | hCoV-19/Switzerland/BE-ETHZ-250119/2020\|EPI_ISL_53 | ···········T········ | ···················· | ···················· | ···················· | ···················· | ···················· |
| H | 6998 | hCoV-19/Switzerland/BS-ETHZ-250121/2020\|EPI_ISL_53 | ···················· | ·A·················· | ···················· | ···················· | ···················· | ···················· |
| H | 6999 | hCoV-19/Switzerland/BL-ETHZ-250123/2020\|EPI_ISL_53 | ···················· | ·A·················· | ···················· | ···················· | ···················· | ···················· |
| H | 7000 | hCoV-19/Switzerland/BS-ETHZ-250124/2020\|EPI_ISL_53 | ···················· | ···················· | ···················· | ···················· | ···················· | ···················· |
| H | 7001 | hCoV-19/Switzerland/BL-ETHZ-250125/2020\|EPI_ISL_53 | ···················· | ·A·················· | ···················· | ···················· | ···················· | ···················· |
| H | 7002 | hCoV-19/Switzerland/BL-ETHZ-250126/2020\|EPI_ISL_53 | ···················· | ···················· | ···················· | ···················· | ···················· | ···················· |
| H | 7003 | hCoV-19/Switzerland/BS-ETHZ-250134/2020\|EPI_ISL_53 | ···················· | ···················· | ···················· | ···················· | ···················· | ···················· |
| H | 7004 | hCoV-19/Switzerland/GR-ETHZ-250135/2020\|EPI_ISL_53 | ···················· | ···················· | ···················· | ···················· | ···················· | ···················· |
| H | 7005 | hCoV-19/Switzerland/ZH-ETHZ-250138/2020\|EPI_ISL_53 | ···················· | ···················· | ···················· | ···················· | ···················· | ···················· |
| H | 7006 | hCoV-19/Switzerland/SO-ETHZ-250139/2020\|EPI_ISL_53 | ···················· | ···················· | ···················· | ···················· | ···················· | ···················· |
| H | 7007 | hCoV-19/Switzerland/ZH-ETHZ-250141/2020\|EPI_ISL_53 | ···················· | ···················· | ···················· | ···················· | ···················· | ···················· |
| H | 7008 | hCoV-19/Switzerland/VD-ETHZ-250143/2020\|EPI_ISL_53 | ···················· | ···················· | ···················· | ···················· | ···················· | ···················· |
| H | 7009 | hCoV-19/Switzerland/VD-ETHZ-250145/2020\|EPI_ISL_53 | ···················· | ···················· | ···················· | ···················· | ···················· | ···················· |
| H | 7010 | hCoV-19/USA/AR-CDC-0837/2020\|EPI_ISL_535650\|2020-0 | ···················· | ···················· | ···················· | ···················· | ···················· | ···················· |
| H | 7011 | hCoV-19/USA/AR-CDC-0838/2020\|EPI_ISL_535651\|2020-0 | ···················· | ···················· | ···················· | ···················· | ···················· | ···················· |
| H | 7012 | hCoV-19/USA/AR-CDC-0840/2020\|EPI_ISL_535652\|2020-0 | ···················· | ···················· | ···················· | ···················· | ···················· | ···················· |
| H | 7013 | hCoV-19/USA/AR-CDC-0843/2020\|EPI_ISL_535653\|2020-0 | ···················· | ···················· | ···················· | ···················· | ···················· | ···················· |
| H | 7014 | hCoV-19/USA/AR-CDC-0848/2020\|EPI_ISL_535654\|2020-0 | ···················· | ···················· | ···················· | ···················· | ···················· | ···················· |
| H | 7015 | hCoV-19/USA/AR-CDC-0852/2020\|EPI_ISL_535655\|2020-0 | ···················· | ···················· | ···················· | ···················· | ···················· | ···················· |
| H | 7016 | hCoV-19/USA/AR-CDC-0863/2020\|EPI_ISL_535656\|2020-0 | ···················· | ···················· | ···················· | ···················· | ···················· | ···················· |

| | | | | | | | | |
|---|---|---|---|---|---|---|---|---|
| H | 7017 | hCoV-19/USA/AR-CDC-0865/2020\|EPI_ISL_535657\|2020-0 | ···················· | ···················· | ····················· | ···················· | ···················· | ···················· |
| H | 7018 | hCoV-19/England/QEUH-96A3B7/2020\|EPI_ISL_536904\|20 | ···················· | ···················· | ····················· | ···················· | ···················· | ···················· |
| H | 7019 | hCoV-19/England/QEUH-964836/2020\|EPI_ISL_536905\|20 | ···················· | ···················· | ····················· | ···················· | ···················· | ···················· |
| H | 7020 | hCoV-19/Northern Ireland/QEUH-963FD8/2020\|EPI_ISL_ | ···················· | ···················· | ····················· | ···················· | ···················· | ···················· |
| H | 7021 | hCoV-19/England/QEUH-9645F3/2020\|EPI_ISL_536907\|20 | ···················· | ···················· | ····················· | ···················· | ···················· | ···················· |
| H | 7022 | hCoV-19/England/QEUH-964265/2020\|EPI_ISL_536908\|20 | ···················· | ···················· | ····················· | ···················· | ···················· | ···················· |
| H | 7023 | hCoV-19/England/QEUH-96A399/2020\|EPI_ISL_536909\|20 | ···················· | ···················· | ····················· | ···················· | ···················· | ···················· |
| H | 7024 | hCoV-19/England/QEUH-964863/2020\|EPI_ISL_536910\|20 | ···················· | ···················· | ····················· | ···················· | ···················· | ···················· |
| H | 7025 | hCoV-19/Northern Ireland/QEUH-9641A4/2020\|EPI_ISL_ | ···················· | ···················· | ····················· | ···················· | ···················· | ···················· |
| H | 7026 | hCoV-19/Northern Ireland/QEUH-9642FC/2020\|EPI_ISL_ | ···················· | ···················· | ····················· | ···················· | ···················· | ···················· |
| H | 7027 | hCoV-19/England/QEUH-964599/2020\|EPI_ISL_536913\|20 | ···················· | ···················· | ····················· | ···················· | ···················· | ···················· |
| H | 7028 | hCoV-19/England/QEUH-96470C/2020\|EPI_ISL_536914\|20 | ···················· | ···················· | ····················· | ···················· | ···················· | ···················· |
| H | 7029 | hCoV-19/England/QEUH-964274/2020\|EPI_ISL_536915\|20 | ···················· | ···················· | ····················· | ···················· | ···················· | ···················· |
| H | 7030 | hCoV-19/England/QEUH-964238/2020\|EPI_ISL_536917\|20 | ···················· | ···················· | ····················· | ···················· | ···················· | ···················· |
| H | 7031 | hCoV-19/Northern Ireland/QEUH-96511C/2020\|EPI_ISL_ | ···················· | ···················· | ····················· | ···················· | ···················· | ···················· |
| H | 7032 | hCoV-19/England/QEUH-96A29C/2020\|EPI_ISL_536919\|20 | ···················· | ···················· | ····················· | ···················· | ···················· | ···················· |
| H | 7033 | hCoV-19/Northern Ireland/QEUH-964405/2020\|EPI_ISL_ | ···················· | ···················· | ····················· | ···················· | ···················· | ···················· |
| H | 7034 | hCoV-19/Northern Ireland/QEUH-964159/2020\|EPI_ISL_ | ···················· | ···················· | ····················· | ···················· | ···················· | ···················· |
| H | 7035 | hCoV-19/England/QEUH-964845/2020\|EPI_ISL_536922\|20 | ···················· | ···················· | ····················· | ···················· | ···················· | ···················· |
| H | 7036 | hCoV-19/England/QEUH-9646F0/2020\|EPI_ISL_536923\|20 | ···················· | ···················· | ····················· | ···················· | ···················· | ···················· |
| H | 7037 | hCoV-19/Wales/QEUH-96A180/2020\|EPI_ISL_536924\|2020 | ···················· | ···················· | ····················· | ···················· | ···················· | ···················· |
| H | 7038 | hCoV-19/England/QEUH-9647EE/2020\|EPI_ISL_536925\|20 | ···················· | ···················· | ····················· | ···················· | ···················· | ···················· |
| H | 7039 | hCoV-19/Northern Ireland/QEUH-963F9C/2020\|EPI_ISL_ | ···················· | ···················· | ····················· | ···················· | ···················· | ···················· |
| H | 7040 | hCoV-19/England/QEUH-96439F/2020\|EPI_ISL_536927\|20 | ···················· | ···················· | ····················· | ···················· | ···················· | ···················· |
| H | 7041 | hCoV-19/England/QEUH-96A0A1/2020\|EPI_ISL_536928\|20 | ···················· | ···················· | ····················· | ···················· | ···················· | ···················· |
| H | 7042 | hCoV-19/England/QEUH-96A117/2020\|EPI_ISL_536929\|20 | ···················· | ·A·················· | ····················· | ···················· | ···················· | ··A················· |
| H | 7043 | hCoV-19/Northern Ireland/QEUH-96410E/2020\|EPI_ISL_ | ···················· | ···················· | ····················· | ···················· | ···················· | ···················· |
| H | 7044 | hCoV-19/England/QEUH-96A1F9/2020\|EPI_ISL_536932\|20 | ···················· | ···················· | ····················· | ···················· | ···················· | ···················· |
| H | 7045 | hCoV-19/Northern Ireland/QEUH-9650C4/2020\|EPI_ISL_ | ···················· | ···················· | ····················· | ···················· | ···················· | ···················· |
| H | 7046 | hCoV-19/England/QEUH-9642A1/2020\|EPI_ISL_536934\|20 | ···················· | ···················· | ····················· | ···················· | ···················· | ···················· |
| H | 7047 | hCoV-19/Northern Ireland/QEUH-96411D/2020\|EPI_ISL_ | ···················· | ···················· | ····················· | ···················· | ···················· | ···················· |
| H | 7048 | hCoV-19/England/QEUH-96A2AB/2020\|EPI_ISL_536937\|20 | ···················· | ···················· | ····················· | ···················· | ···················· | ···················· |
| H | 7049 | hCoV-19/England/QEUH-9648BE/2020\|EPI_ISL_536938\|20 | ···················· | ···················· | ····················· | ···················· | ···················· | ···················· |
| H | 7050 | hCoV-19/England/QEUH-96A162/2020\|EPI_ISL_536939\|20 | ···················· | ···················· | ····················· | ···················· | ···················· | ···················· |
| H | 7051 | hCoV-19/England/QEUH-96A2D8/2020\|EPI_ISL_536940\|20 | ···················· | ···················· | ····················· | ···················· | ···················· | ···················· |
| H | 7052 | hCoV-19/England/QEUH-963AF5/2020\|EPI_ISL_536943\|20 | ···················· | ···················· | ····················· | ···················· | ···················· | ···················· |
| H | 7053 | hCoV-19/England/QEUH-9651A3/2020\|EPI_ISL_536944\|20 | ···················· | ···················· | ····················· | ···················· | ···················· | ···················· |
| H | 7054 | hCoV-19/England/QEUH-96A223/2020\|EPI_ISL_536945\|20 | ···················· | ···················· | ····················· | ···················· | ······T············· | ···················· |
| H | 7055 | hCoV-19/England/QEUH-96A0CF/2020\|EPI_ISL_536947\|20 | ···················· | ···················· | ····················· | ···················· | ···················· | ···················· |
| H | 7056 | hCoV-19/England/QEUH-964450/2020\|EPI_ISL_536948\|20 | ···················· | ···················· | ····················· | ···················· | ···················· | ···················· |
| H | 7057 | hCoV-19/Northern Ireland/QEUH-96402F/2020\|EPI_ISL_ | ···················· | ···················· | ····················· | ···················· | ···················· | ···················· |
| H | 7058 | hCoV-19/England/QEUH-964186/2020\|EPI_ISL_536950\|20 | ···················· | ···················· | ····················· | ···················· | ···················· | ···················· |
| H | 7059 | hCoV-19/England/QEUH-963F23/2020\|EPI_ISL_536952\|20 | ···················· | ···················· | ····················· | ···················· | ···················· | ···················· |
| H | 7060 | hCoV-19/England/QEUH-96A311/2020\|EPI_ISL_536953\|20 | ···················· | ···················· | ····················· | ···················· | ···················· | ···················· |
| H | 7061 | hCoV-19/England/QEUH-9643F9/2020\|EPI_ISL_536954\|20 | ···················· | ···················· | ····················· | ···················· | ···················· | ···················· |
| H | 7062 | hCoV-19/England/QEUH-96A205/2020\|EPI_ISL_536958\|20 | ···················· | ···················· | ····················· | ···················· | ···················· | ···················· |
| H | 7063 | hCoV-19/England/QEUH-96471B/2020\|EPI_ISL_536959\|20 | ···················· | ···················· | ····················· | ···················· | ···················· | ···················· |
| H | 7064 | hCoV-19/England/QEUH-96449C/2020\|EPI_ISL_536960\|20 | ···················· | ···················· | ····················· | ···················· | ···················· | ···················· |
| H | 7065 | hCoV-19/England/QEUH-964784/2020\|EPI_ISL_536961\|20 | ···················· | ···················· | ····················· | ···················· | ···················· | ···················· |
| H | 7066 | hCoV-19/England/QEUH-961972/2020\|EPI_ISL_536962\|20 | ···················· | ···················· | ····················· | ···················· | ···················· | ···················· |
| H | 7067 | hCoV-19/England/QEUH-95E3A7/2020\|EPI_ISL_536963\|20 | ···················· | ···················· | ····················· | ···················· | ···················· | ···················· |
| H | 7068 | hCoV-19/England/QEUH-961408/2020\|EPI_ISL_536964\|20 | ···················· | ···················· | ····················· | ···················· | ···················· | ···················· |
| H | 7069 | hCoV-19/England/QEUH-961E19/2020\|EPI_ISL_536966\|20 | ···················· | ···················· | ····················· | ···················· | ···················· | ···················· |
| H | 7070 | hCoV-19/England/QEUH-9618B1/2020\|EPI_ISL_536967\|20 | ···················· | ···················· | ····················· | ···················· | ···················· | ···················· |
| H | 7071 | hCoV-19/England/QEUH-961866/2020\|EPI_ISL_536968\|20 | ···················· | ···················· | ····················· | ···················· | ···················· | ···················· |
| H | 7072 | hCoV-19/England/QEUH-95E240/2020\|EPI_ISL_536969\|20 | ···················· | ···················· | ····················· | ···················· | ···················· | ···················· |
| H | 7073 | hCoV-19/England/QEUH-95DF0E/2020\|EPI_ISL_536970\|20 | ···················· | ···················· | ····················· | ···················· | ···················· | ···················· |
| H | 7074 | hCoV-19/England/QEUH-95DCF8/2020\|EPI_ISL_536971\|20 | ···················· | ···················· | ····················· | ···················· | ···················· | ···················· |
| H | 7075 | hCoV-19/England/QEUH-9613CF/2020\|EPI_ISL_536972\|20 | ···················· | ···················· | ····················· | ···················· | ···················· | ···················· |
| H | 7076 | hCoV-19/England/QEUH-9616C6/2020\|EPI_ISL_536973\|20 | ···················· | ···················· | ····················· | ···················· | ···················· | ···················· |
| H | 7077 | hCoV-19/England/QEUH-95DCDA/2020\|EPI_ISL_536974\|20 | ···················· | ·A·················· | ····················· | ···················· | ···················· | ···················· |
| H | 7078 | hCoV-19/England/QEUH-9619AF/2020\|EPI_ISL_536975\|20 | ···················· | ···················· | ····················· | ···················· | ···················· | ···················· |
| H | 7079 | hCoV-19/England/QEUH-961A7F/2020\|EPI_ISL_536976\|20 | ···················· | ···················· | ····················· | ···················· | ···················· | ···················· |
| H | 7080 | hCoV-19/Northern Ireland/QEUH-961CE2/2020\|EPI_ISL_ | ···················· | ···················· | ····················· | ···················· | ···················· | ···················· |
| H | 7081 | hCoV-19/England/QEUH-961DC1/2020\|EPI_ISL_536978\|20 | ···················· | ···················· | ····················· | ···················· | ···················· | ···················· |
| H | 7082 | hCoV-19/England/QEUH-961A24/2020\|EPI_ISL_536979\|20 | ···················· | ···················· | ····················· | ···················· | ···················· | ···················· |
| H | 7083 | hCoV-19/England/QEUH-961E64/2020\|EPI_ISL_536980\|20 | ···················· | ···················· | ···········T········· | ···················· | ···················· | ···················· |
| H | 7084 | hCoV-19/England/QEUH-961D0D/2020\|EPI_ISL_536981\|20 | ···················· | ···················· | ····················· | ···················· | ···················· | ···················· |
| H | 7085 | hCoV-19/England/QEUH-95DDAA/2020\|EPI_ISL_536982\|20 | ···················· | ·A·················· | ····················· | ···················· | ···················· | ··A················· |
| H | 7086 | hCoV-19/England/QEUH-95DE3E/2020\|EPI_ISL_536983\|20 | ···················· | ·A·················· | ····················· | ···················· | ···················· | ··A················· |
| H | 7087 | hCoV-19/England/QEUH-961426/2020\|EPI_ISL_536984\|20 | ···················· | ···················· | ····················· | ···················· | ···················· | ···················· |
| H | 7088 | hCoV-19/England/QEUH-961787/2020\|EPI_ISL_536985\|20 | ···················· | ·A·················· | ····················· | ···················· | ···················· | ··A················· |
| H | 7089 | hCoV-19/England/QEUH-961893/2020\|EPI_ISL_536986\|20 | ···················· | ·A·················· | ····················· | ···················· | ···················· | ···················· |
| H | 7090 | hCoV-19/England/QEUH-95DD13/2020\|EPI_ISL_536987\|20 | ···················· | ···················· | ····················· | ···················· | ···················· | ···················· |
| H | 7091 | hCoV-19/England/QEUH-9617D2/2020\|EPI_ISL_536988\|20 | ···················· | ···················· | ····················· | ···················· | ···················· | ···················· |
| H | 7092 | hCoV-19/England/LEED-2A916E/2020\|EPI_ISL_539180\|20 | ···················· | ·A·················· | ····················· | ···················· | ···················· | ···················· |
| H | 7093 | hCoV-19/England/LEED-2A92E3/2020\|EPI_ISL_539183\|20 | ···················· | ···················· | ····················· | ···················· | ···················· | ···················· |
| H | 7094 | hCoV-19/England/LEED-2A8DD4/2020\|EPI_ISL_539184\|20 | ···················· | ···················· | ····················· | ···················· | ···················· | ···················· |
| H | 7095 | hCoV-19/England/LEED-2A8527/2020\|EPI_ISL_539186\|20 | ···················· | ···················· | ····················· | ···················· | ···················· | ···················· |
| H | 7096 | hCoV-19/England/LEED-2A84FD/2020\|EPI_ISL_539187\|20 | ···················· | ···················· | ····················· | ···················· | ···················· | ···················· |

| | | | | | | | |
|---|---|---|---|---|---|---|---|
| H | 7097 hCoV-19/England/LEED-2A9298/2020\|EPI_ISL_539193\|20 | .................... | .................... | ..................... | .................... | .................... | .................... |
| H | 7098 hCoV-19/England/LEED-2A842A/2020\|EPI_ISL_539194\|20 | .................... | .................... | ..................... | .................... | .................... | .................... |
| H | 7099 hCoV-19/England/LEED-2A84EE/2020\|EPI_ISL_539195\|20 | .................... | .................... | ..................... | .................... | .................... | .................... |
| H | 7100 hCoV-19/England/LEED-2A8C40/2020\|EPI_ISL_539196\|20 | .................... | .................... | ..................... | .................... | .................... | .................... |
| H | 7101 hCoV-19/England/LEED-2A93C2/2020\|EPI_ISL_539198\|20 | .................... | .................... | ..................... | .................... | .................... | .................... |
| H | 7102 hCoV-19/England/LEED-2A92B6/2020\|EPI_ISL_539199\|20 | .................... | .................... | ..................... | .................... | .................... | .................... |
| H | 7103 hCoV-19/England/QEUH-9A4436/2020\|EPI_ISL_540168\|20 | .................... | .................... | ..................... | .................... | .................... | .................... |
| H | 7104 hCoV-19/England/QEUH-9A459D/2020\|EPI_ISL_540169\|20 | .................... | .................... | ..................... | .................... | .................... | .................... |
| H | 7105 hCoV-19/England/QEUH-9A466D/2020\|EPI_ISL_540170\|20 | .................... | .................... | ..................... | .................... | .................... | .................... |
| H | 7106 hCoV-19/England/QEUH-9A4296/2020\|EPI_ISL_540171\|20 | .................... | .A.................. | ..................... | .................... | .................... | .................... |
| H | 7107 hCoV-19/England/QEUH-9A468B/2020\|EPI_ISL_540172\|20 | .................... | .................... | ..................... | .................... | .................... | .................... |
| H | 7108 hCoV-19/England/QEUH-9A4481/2020\|EPI_ISL_540173\|20 | .................... | .................... | ..................... | .................... | .................... | .................... |
| H | 7109 hCoV-19/England/QEUH-9A418A/2020\|EPI_ISL_540174\|20 | .................... | .................... | ..................... | .................... | .................... | .................... |
| H | 7110 hCoV-19/England/QEUH-9A4524/2020\|EPI_ISL_540175\|20 | .................... | .................... | ..................... | .................... | .................... | .................... |
| H | 7111 hCoV-19/England/QEUH-9A41B7/2020\|EPI_ISL_540176\|20 | .................... | .................... | ..................... | .................... | .................... | .................... |
| H | 7112 hCoV-19/England/QEUH-9A4418/2020\|EPI_ISL_540177\|20 | .................... | .................... | ..................... | .................... | .................... | .................... |
| H | 7113 hCoV-19/England/QEUH-9A465E/2020\|EPI_ISL_540178\|20 | .................... | .................... | ..................... | .................... | .................... | .................... |
| H | 7114 hCoV-19/England/QEUH-9A46A9/2020\|EPI_ISL_540179\|20 | .................... | .................... | ..................... | .................... | .................... | .................... |
| H | 7115 hCoV-19/England/QEUH-9A475B/2020\|EPI_ISL_540180\|20 | .................... | .................... | ..................... | .................... | .................... | .................... |
| H | 7116 hCoV-19/England/QEUH-9A4463/2020\|EPI_ISL_540181\|20 | .................... | .A.......A.......... | ..................... | .................... | .................... | .................... |
| H | 7117 hCoV-19/England/QEUH-9A4560/2020\|EPI_ISL_540182\|20 | .................... | .................... | ...........T......... | .................... | .................... | .................... |
| H | 7118 hCoV-19/England/QEUH-9A4445/2020\|EPI_ISL_540183\|20 | .................... | .................... | ..................... | .................... | .................... | .................... |
| H | 7119 hCoV-19/England/QEUH-9A4454/2020\|EPI_ISL_540185\|20 | .................... | .A.................. | ..................... | .................... | .................... | .................... |
| H | 7120 hCoV-19/England/QEUH-9A4348/2020\|EPI_ISL_540186\|20 | .................... | .................... | ..................... | .................... | .................... | .................... |
| H | 7121 hCoV-19/England/QEUH-9A4472/2020\|EPI_ISL_540187\|20 | .................... | .................... | ..................... | .................... | .................... | .................... |
| H | 7122 hCoV-19/England/QEUH-9A44FA/2020\|EPI_ISL_540188\|20 | .................... | .................... | ..................... | .................... | .................... | .................... |
| H | 7123 hCoV-19/England/QEUH-9A4111/2020\|EPI_ISL_540189\|20 | .................... | .................... | ..................... | .................... | .................... | .................... |
| H | 7124 hCoV-19/England/QEUH-9A44AF/2020\|EPI_ISL_540191\|20 | .................... | .A.................. | ..................... | .................... | .................... | .................... |
| H | 7125 hCoV-19/England/QEUH-9A4542/2020\|EPI_ISL_540192\|20 | .................... | .................... | ..................... | .................... | .................... | .................... |
| H | 7126 hCoV-19/England/QEUH-9A41E4/2020\|EPI_ISL_540193\|20 | .................... | .................... | ..................... | .................... | .................... | .................... |
| H | 7127 hCoV-19/England/QEUH-9A4409/2020\|EPI_ISL_540194\|20 | .................... | .................... | ..................... | .................... | .................... | .................... |
| H | 7128 hCoV-19/England/QEUH-9A4287/2020\|EPI_ISL_540195\|20 | .................... | .................... | ..................... | .................... | .................... | .................... |
| H | 7129 hCoV-19/England/QEUH-9A416C/2020\|EPI_ISL_540196\|20 | .................... | .................... | ..................... | .................... | .................... | .................... |
| H | 7130 hCoV-19/England/QEUH-9A4357/2020\|EPI_ISL_540197\|20 | .................... | .................... | ..................... | .................... | .................... | .................... |
| H | 7131 hCoV-19/England/QEUH-9A4630/2020\|EPI_ISL_540198\|20 | .................... | .A.......A.......... | ..................... | .................... | .................... | .................... |
| H | 7132 hCoV-19/England/QEUH-9A3FAF/2020\|EPI_ISL_540199\|20 | .................... | .................... | ..................... | .................... | .................... | .................... |
| H | 7133 hCoV-19/England/QEUH-9A40F6/2020\|EPI_ISL_540201\|20 | .................... | .................... | ..................... | .................... | .................... | .................... |
| H | 7134 hCoV-19/England/QEUH-9A414E/2020\|EPI_ISL_540202\|20 | .................... | .................... | ..................... | .................... | .................... | .................... |
| H | 7135 hCoV-19/England/QEUH-9A45D9/2020\|EPI_ISL_540203\|20 | .................... | .................... | ..................... | .................... | ....T............... | .................... |
| H | 7136 hCoV-19/England/QEUH-9A44BE/2020\|EPI_ISL_540204\|20 | .................... | .................... | ..................... | .................... | .................... | .................... |
| H | 7137 hCoV-19/England/QEUH-9A408D/2020\|EPI_ISL_540205\|20 | .................... | .................... | ..................... | .................... | .................... | .................... |
| H | 7138 hCoV-19/England/QEUH-9A43EE/2020\|EPI_ISL_540206\|20 | .................... | .................... | ..................... | .................... | .................... | .................... |
| H | 7139 hCoV-19/England/QEUH-9A4621/2020\|EPI_ISL_540207\|20 | .................... | .................... | ..................... | .................... | .................... | .................... |
| H | 7140 hCoV-19/England/QEUH-9A3FEB/2020\|EPI_ISL_540208\|20 | .................... | .................... | ..................... | .................... | .................... | .................... |
| H | 7141 hCoV-19/England/QEUH-9A40C9/2020\|EPI_ISL_540209\|20 | .................... | .................... | ..................... | .................... | .................... | .................... |
| H | 7142 hCoV-19/England/QEUH-9A457F/2020\|EPI_ISL_540210\|20 | .................... | .................... | ...........T......... | .................... | .................... | .................... |
| H | 7143 hCoV-19/England/QEUH-9A3FDC/2020\|EPI_ISL_540211\|20 | .................... | .................... | ..................... | .................... | .................... | .................... |
| H | 7144 hCoV-19/Switzerland/VD-ETHZ-270175/2020\|EPI_ISL_54 | .................... | .................... | ..................... | .................... | .................... | .................... |
| H | 7145 hCoV-19/Switzerland/ZH-ETHZ-270185/2020\|EPI_ISL_54 | .................... | .................... | ..................... | .................... | .................... | .................... |
| H | 7146 hCoV-19/Switzerland/ZH-ETHZ-270186/2020\|EPI_ISL_54 | .................... | .................... | ..................... | .................... | .................... | .................... |
| H | 7147 hCoV-19/Switzerland/ZH-ETHZ-270190/2020\|EPI_ISL_54 | .................... | .................... | ..................... | .................... | .................... | .................... |
| H | 7148 hCoV-19/Switzerland/ZH-ETHZ-270193/2020\|EPI_ISL_54 | .................... | .................... | ..................... | .................... | .................... | .................... |
| H | 7149 hCoV-19/Switzerland/ZH-ETHZ-270198/2020\|EPI_ISL_54 | .................... | .................... | ..................... | .................... | ......K............. | .................... |
| H | 7150 hCoV-19/Switzerland/ZH-ETHZ-270206/2020\|EPI_ISL_54 | .................... | .................... | ..................... | .................... | .................... | .................... |
| H | 7151 hCoV-19/Switzerland/ZH-ETHZ-270207/2020\|EPI_ISL_54 | .................... | .................... | ..................... | .................... | .................... | .................... |
| H | 7152 hCoV-19/USA/IL-UW-1439/2020\|EPI_ISL_541540\|2020-09 | .................... | .A.................. | ..................... | .................... | .................... | .................... |
| H | 7153 hCoV-19/USA/IL-UW-1481/2020\|EPI_ISL_541541\|2020-09 | .................... | .................... | ..................... | .................... | .................... | .................... |
| H | 7154 hCoV-19/USA/IL-UW-1483/2020\|EPI_ISL_541542\|2020-09 | .................... | .................... | ..................... | .................... | .................... | .................... |
| H | 7155 hCoV-19/USA/TX-HMH-1820/2020\|EPI_ISL_542863\|2020-0 | .................... | .................... | ..................... | .................... | .................... | .................... |
| H | 7156 hCoV-19/USA/TX-HMH-1822/2020\|EPI_ISL_542864\|2020-0 | .................... | ......A............. | ..................... | .................... | .................... | .................... |
| H | 7157 hCoV-19/USA/TX-HMH-1824/2020\|EPI_ISL_542865\|2020-0 | .................... | .................... | ..................... | .................... | .................... | .................... |
| H | 7158 hCoV-19/USA/TX-HMH-1825/2020\|EPI_ISL_542866\|2020-0 | .................... | .................... | ..................... | .................... | .................... | .................... |
| H | 7159 hCoV-19/USA/TX-HMH-1828/2020\|EPI_ISL_542868\|2020-0 | .................... | ......A............. | ..................... | .................... | .................... | .................... |
| H | 7160 hCoV-19/USA/TX-HMH-1830/2020\|EPI_ISL_542869\|2020-0 | .................... | .................... | ..................... | .................... | .................... | .................... |
| H | 7161 hCoV-19/USA/TX-HMH-1832/2020\|EPI_ISL_542871\|2020-0 | .................... | .................... | ..................... | .................... | .................... | .................... |
| H | 7162 hCoV-19/USA/TX-HMH-1835/2020\|EPI_ISL_542872\|2020-0 | .................... | .................... | ..................... | .................... | .................... | .................... |
| H | 7163 hCoV-19/USA/TX-HMH-1836/2020\|EPI_ISL_542873\|2020-0 | .................... | .................... | ..................... | .................... | .................... | .................... |
| H | 7164 hCoV-19/Pakistan/UN-UVAS-Lahore-I/2020\|EPI_ISL_548 | .................... | .................... | ..................... | .................... | .................... | .................... |
| H | 7165 hCoV-19/Pakistan/UN-UVAS-Lahore-II/2020\|EPI_ISL_54 | .................... | .................... | ..................... | .................... | .................... | .................... |
| H | 7166 hCoV-19/Pakistan/UN-UVAS-Lahore-III/2020\|EPI_ISL_5 | .................... | .................... | ..................... | .................... | .................... | .................... |
| H | 7167 hCoV-19/Pakistan/UN-UVAS-Lahore-IV/2020\|EPI_ISL_54 | .................... | .................... | ..................... | .................... | .................... | .................... |
| H | 7168 hCoV-19/Pakistan/UN-UVAS-Sialkot/2020\|EPI_ISL_5489 | .................... | .................... | ..................... | .................... | .................... | .................... |
| H | 7169 hCoV-19/England/QEUH-96D132/2020\|EPI_ISL_530801\|20 | .................... | .A.................. | ..................... | .................... | .................... | .................... |
| H | 7170 hCoV-19/England/QEUH-96A8F4/2020\|EPI_ISL_530802\|20 | .................... | .................... | ..................... | .................... | .................... | .................... |
| H | 7171 hCoV-19/England/QEUH-96C643/2020\|EPI_ISL_530803\|20 | .................... | .................... | ..................... | .................... | .................... | .................... |
| H | 7172 hCoV-19/England/QEUH-96C3F1/2020\|EPI_ISL_530804\|20 | .................... | .................... | ..................... | .................... | .................... | .................... |
| H | 7173 hCoV-19/England/QEUH-96C6E9/2020\|EPI_ISL_530805\|20 | .................... | .................... | ..................... | .................... | .................... | .................... |
| H | 7174 hCoV-19/England/QEUH-96C537/2020\|EPI_ISL_530806\|20 | .................... | .................... | ..................... | .................... | .................... | .................... |
| H | 7175 hCoV-19/England/QEUH-96A8B8/2020\|EPI_ISL_530807\|20 | .................... | .................... | ..................... | .................... | .................... | .................... |
| H | 7176 hCoV-19/England/QEUH-969D23/2020\|EPI_ISL_530808\|20 | .................... | .................... | ..................... | .................... | .................... | .................... |

| | | | | | | | | |
|---|---|---|---|---|---|---|---|---|
| H | 7177 | hCoV-19/Northern Ireland/QEUH-969DBA/2020\|EPI_ISL_ | ···················· | ···················· | ···················· | ···················· | ···················· | ···················· |
| H | 7178 | hCoV-19/Northern Ireland/QEUH-96C133/2020\|EPI_ISL_ | ···················· | ···················· | ···················· | ···················· | ···················· | ···················· |
| H | 7179 | hCoV-19/England/QEUH-96C17F/2020\|EPI_ISL_530811\|20 | ···················· | ···················· | ···················· | ···················· | ···················· | ···················· |
| H | 7180 | hCoV-19/England/QEUH-96C1BB/2020\|EPI_ISL_530812\|20 | ···················· | ···················· | ···················· | ···················· | ···················· | ···················· |
| H | 7181 | hCoV-19/England/QEUH-969C9F/2020\|EPI_ISL_530813\|20 | ···················· | ···················· | ···················· | ···················· | ···················· | ···················· |
| H | 7182 | hCoV-19/England/QEUH-96C1E8/2020\|EPI_ISL_530814\|20 | ···················· | ···················· | ···················· | ···················· | ···················· | ···················· |
| H | 7183 | hCoV-19/England/QEUH-96C4FE/2020\|EPI_ISL_530815\|20 | ···················· | ···················· | ···················· | ···················· | ···················· | ···················· |
| H | 7184 | hCoV-19/England/QEUH-96C68F/2020\|EPI_ISL_530816\|20 | ···················· | ···················· | ···················· | ···················· | ···················· | ···················· |
| H | 7185 | hCoV-19/England/QEUH-969F3C/2020\|EPI_ISL_530817\|20 | ···················· | ···················· | ···················· | ···················· | ···················· | ···················· |
| H | 7186 | hCoV-19/England/QEUH-96A01A/2020\|EPI_ISL_530818\|20 | ···················· | ···················· | ···················· | ···················· | ···················· | ···················· |
| H | 7187 | hCoV-19/England/QEUH-96A047/2020\|EPI_ISL_530819\|20 | ···················· | ···················· | ···················· | ···················· | ···················· | ···················· |
| H | 7188 | hCoV-19/England/QEUH-969F69/2020\|EPI_ISL_530820\|20 | ···················· | ···················· | ···················· | ···················· | ···················· | ···················· |
| H | 7189 | hCoV-19/Northern Ireland/QEUH-96C0FA/2020\|EPI_ISL_ | ···················· | ···················· | ···················· | ···················· | ···················· | ···················· |
| H | 7190 | hCoV-19/Northern Ireland/QEUH-96C142/2020\|EPI_ISL_ | ···················· | ···················· | ···················· | ···················· | ···················· | ···················· |
| H | 7191 | hCoV-19/Northern Ireland/QEUH-969D7E/2020\|EPI_ISL_ | ···················· | ···················· | ···················· | ···················· | ···················· | ···················· |
| H | 7192 | hCoV-19/England/QEUH-96C397/2020\|EPI_ISL_530824\|20 | ···················· | ···················· | ···················· | ···················· | ···················· | ···················· |
| H | 7193 | hCoV-19/England/QEUH-969D41/2020\|EPI_ISL_530825\|20 | ···················· | ···················· | ···················· | ···················· | ···················· | ···················· |
| H | 7194 | hCoV-19/Scotland/QEUH-96C0BE/2020\|EPI_ISL_530826\|2 | ···················· | ···················· | ···················· | ···················· | ···················· | ···················· |
| H | 7195 | hCoV-19/England/QEUH-969CCC/2020\|EPI_ISL_530827\|20 | ···················· | ···················· | ···················· | ···················· | ···················· | ···················· |
| H | 7196 | hCoV-19/Northern Ireland/QEUH-96C41C/2020\|EPI_ISL_ | ···················· | ···················· | ···················· | ···················· | ···················· | ···················· |
| H | 7197 | hCoV-19/England/QEUH-96C485/2020\|EPI_ISL_530829\|20 | ···················· | ···················· | ···················· | ···················· | ···················· | ···················· |
| H | 7198 | hCoV-19/Northern Ireland/QEUH-969EA8/2020\|EPI_ISL_ | ···················· | ···················· | ···················· | ···················· | ···················· | ···················· |
| H | 7199 | hCoV-19/England/QEUH-96C19D/2020\|EPI_ISL_530831\|20 | ···················· | ···················· | ···················· | ···················· | ···················· | ···················· |
| H | 7200 | hCoV-19/England/QEUH-96A108/2020\|EPI_ISL_530832\|20 | ···················· | ···················· | ···················· | ···················· | ···················· | ···················· |
| H | 7201 | hCoV-19/Northern Ireland/QEUH-96C4B2/2020\|EPI_ISL_ | ···················· | ···················· | ···················· | ···················· | ···················· | ···················· |
| H | 7202 | hCoV-19/England/QEUH-969E99/2020\|EPI_ISL_530834\|20 | ···················· | ···················· | ···················· | ···················· | ···················· | ···················· |
| H | 7203 | hCoV-19/Northern Ireland/QEUH-969C53/2020\|EPI_ISL_ | ···················· | ···················· | ···················· | ···················· | ···················· | ···················· |
| H | 7204 | hCoV-19/Scotland/QEUH-96C212/2020\|EPI_ISL_530836\|2 | ···················· | ···················· | ···················· | ···················· | ···················· | ···················· |
| H | 7205 | hCoV-19/England/QEUH-96C704/2020\|EPI_ISL_530837\|20 | ···················· | ···················· | ···················· | ···················· | ···················· | ···················· |
| H | 7206 | hCoV-19/England/QEUH-96A89A/2020\|EPI_ISL_530838\|20 | ···················· | ·A·················· | ···················· | ···················· | ···················· | ···················· |
| H | 7207 | hCoV-19/England/QEUH-969BED/2020\|EPI_ISL_530839\|20 | ···················· | ···················· | ···················· | ···················· | ···················· | ···················· |
| H | 7208 | hCoV-19/England/QEUH-96A00B/2020\|EPI_ISL_530840\|20 | ···················· | ···················· | ···················· | ···················· | ···················· | ···················· |
| H | 7209 | hCoV-19/England/QEUH-969C08/2020\|EPI_ISL_530841\|20 | ···················· | ·A·················· | ···················· | ···················· | ···················· | ··A················· |
| H | 7210 | hCoV-19/England/QEUH-969CF9/2020\|EPI_ISL_530842\|20 | ···················· | ·A·················· | ···················· | ···················· | ···················· | ··A················· |
| H | 7211 | hCoV-19/England/QEUH-96A083/2020\|EPI_ISL_530843\|20 | ···················· | ···················· | ···················· | ···················· | ···················· | ···················· |
| H | 7212 | hCoV-19/England/QEUH-969CDB/2020\|EPI_ISL_530844\|20 | ···················· | ···················· | ···················· | ···················· | ···················· | ···················· |
| H | 7213 | hCoV-19/Scotland/QEUH-96C458/2020\|EPI_ISL_530845\|2 | ···················· | ···················· | ···················· | ···················· | ···················· | ···················· |
| H | 7214 | hCoV-19/England/QEUH-96C32E/2020\|EPI_ISL_530848\|20 | ···················· | ···················· | ···················· | ···················· | ···················· | ···················· |
| H | 7215 | hCoV-19/England/QEUH-96C1CA/2020\|EPI_ISL_530849\|20 | ···················· | ···················· | ···················· | ···················· | ···················· | ···················· |
| H | 7216 | hCoV-19/England/QEUH-96C625/2020\|EPI_ISL_530850\|20 | ···················· | ···················· | ···················· | ···················· | ···················· | ···················· |
| H | 7217 | hCoV-19/England/QEUH-96C3C4/2020\|EPI_ISL_530851\|20 | ···················· | ···················· | ···················· | ···················· | ···················· | ···················· |
| H | 7218 | hCoV-19/England/QEUH-969C35/2020\|EPI_ISL_530852\|20 | ···················· | ···················· | ···················· | ···················· | ···················· | ···················· |
| H | 7219 | hCoV-19/England/QEUH-96C494/2020\|EPI_ISL_530854\|20 | ···················· | ···················· | ···················· | ···················· | ···················· | ···················· |
| H | 7220 | hCoV-19/Northern Ireland/QEUH-969D6F/2020\|EPI_ISL_ | ···················· | ···················· | ···················· | ···················· | ···················· | ···················· |
| H | 7221 | hCoV-19/England/QEUH-96C2E5/2020\|EPI_ISL_530858\|20 | ···················· | ···················· | ···················· | ···················· | ···················· | ···················· |
| H | 7222 | hCoV-19/England/QEUH-969C71/2020\|EPI_ISL_530859\|20 | ···················· | ···················· | ···················· | ···················· | ···················· | ···················· |
| H | 7223 | hCoV-19/England/QEUH-969ED5/2020\|EPI_ISL_530896\|20 | ···················· | ···················· | ···················· | ···················· | ···················· | ···················· |
| H | 7224 | hCoV-19/England/QEUH-96C34C/2020\|EPI_ISL_530897\|20 | ···················· | ···················· | ···················· | ···················· | ···················· | ···················· |
| H | 7225 | hCoV-19/England/QEUH-78A76C/2020\|EPI_ISL_533080\|20 | ···················· | ···················· | ···················· | ···················· | ···················· | ···················· |
| H | 7226 | hCoV-19/England/QEUH-79EE24/2020\|EPI_ISL_533081\|20 | ···················· | ···················· | ···················· | ···················· | ···················· | ···················· |
| H | 7227 | hCoV-19/Northern Ireland/QEUH-77DAAF/2020\|EPI_ISL_ | ···················· | ···················· | ···················· | ···················· | ···················· | ···················· |
| H | 7228 | hCoV-19/England/QEUH-784B1B/2020\|EPI_ISL_533083\|20 | ···················· | ···················· | ···················· | ···················· | ···················· | ···················· |
| H | 7229 | hCoV-19/England/QEUH-79F1DF/2020\|EPI_ISL_533085\|20 | ···················· | ·A·················· | ···················· | ···················· | ···················· | ···················· |
| H | 7230 | hCoV-19/England/QEUH-79A1E3/2020\|EPI_ISL_533088\|20 | ···················· | ···················· | ···················· | ···················· | ···················· | ···················· |
| H | 7231 | hCoV-19/England/QEUH-79094F/2020\|EPI_ISL_533090\|20 | ···················· | ···················· | ···················· | ···················· | ···················· | ···················· |
| H | 7232 | hCoV-19/England/QEUH-78C630/2020\|EPI_ISL_533092\|20 | ···················· | ···················· | ···················· | ···················· | ···················· | ···················· |
| H | 7233 | hCoV-19/England/QEUH-79ED27/2020\|EPI_ISL_533093\|20 | ···················· | ·A·················· | ···················· | ···················· | ···················· | ···················· |
| H | 7234 | hCoV-19/England/QEUH-797D6A/2020\|EPI_ISL_533095\|20 | ···················· | ···················· | ···················· | ···················· | ···················· | ···················· |
| H | 7235 | hCoV-19/England/QEUH-794B09/2020\|EPI_ISL_533096\|20 | ···················· | ···················· | ···················· | ···················· | ···················· | ···················· |
| H | 7236 | hCoV-19/Northern Ireland/QEUH-77BC14/2020\|EPI_ISL_ | ···················· | ···················· | ···················· | ···················· | ···················· | ···················· |
| H | 7237 | hCoV-19/England/QEUH-7A5B7E/2020\|EPI_ISL_533099\|20 | ···················· | ···················· | ···················· | ···················· | ···················· | ···················· |
| H | 7238 | hCoV-19/Spain/VC-IBV-98003441/2020\|EPI_ISL_539251\| | ···················· | ···················· | ···················· | ···················· | ···················· | ···················· |
| H | 7239 | hCoV-19/Spain/VC-IBV-98003443/2020\|EPI_ISL_539252\| | ···················· | ···················· | ···················· | ···················· | ···················· | ···················· |
| H | 7240 | hCoV-19/Spain/VC-IBV-98003444/2020\|EPI_ISL_539253\| | ···················· | ···················· | ···················· | ···················· | ···················· | ···················· |
| H | 7241 | hCoV-19/Spain/VC-IBV-98003445/2020\|EPI_ISL_539254\| | ···················· | ···················· | ···················· | ···················· | ···················· | ···················· |
| H | 7242 | hCoV-19/Spain/VC-IBV-98003446/2020\|EPI_ISL_539255\| | ···················· | ···················· | ···················· | ···················· | ···················· | ···················· |
| H | 7245 | hCoV-19/Spain/VC-IBV-98003451/2020\|EPI_ISL_539258\| | ···················· | ···················· | ···················· | ···················· | ·NN················· | ···················· |
| H | 7246 | hCoV-19/Spain/VC-IBV-98003489/2020\|EPI_ISL_539281\| | ···················· | ···················· | ···················· | ···················· | ···················· | ···················· |
| H | 7247 | hCoV-19/Spain/VC-IBV-98003490/2020\|EPI_ISL_539282\| | ···················· | ···················· | ···················· | ···················· | ···················· | ···················· |
| H | 7248 | hCoV-19/Spain/VC-IBV-98003492/2020\|EPI_ISL_539283\| | ···················· | ···················· | ···················· | ···················· | ···················· | ···················· |
| H | 7249 | hCoV-19/Spain/VC-IBV-98003494/2020\|EPI_ISL_539284\| | ···················· | ···················· | ···················· | ···················· | ···················· | ···················· |
| H | 7250 | hCoV-19/Spain/VC-IBV-98003495/2020\|EPI_ISL_539285\| | ···················· | ···················· | ···················· | ···················· | ···················· | ···················· |
| H | 7253 | hCoV-19/England/QEUH-9A43B1/2020\|EPI_ISL_540212\|20 | ···················· | ···················· | ···················· | ···················· | ···················· | ···················· |
| H | 7254 | hCoV-19/England/QEUH-9A4102/2020\|EPI_ISL_540213\|20 | ···················· | ···················· | ···················· | ···················· | ···················· | ···················· |
| H | 7255 | hCoV-19/England/QEUH-9A3FFA/2020\|EPI_ISL_540214\|20 | ···················· | ···················· | ···················· | ···················· | ···················· | ···················· |
| H | 7256 | hCoV-19/England/QEUH-9A4199/2020\|EPI_ISL_540215\|20 | ···················· | ···················· | ···················· | ···················· | ···················· | ···················· |
| H | 7257 | hCoV-19/England/QEUH-9A469A/2020\|EPI_ISL_540216\|20 | ···················· | ···················· | ···················· | ···················· | ···················· | ···················· |
| H | 7258 | hCoV-19/Wales/QEUH-9A4506/2020\|EPI_ISL_540217\|2020 | ···················· | ···················· | ···················· | ···················· | ···················· | ···················· |
| H | 7259 | hCoV-19/England/QEUH-9A4515/2020\|EPI_ISL_540218\|20 | ···················· | ···················· | ···················· | ···················· | ···T················ | ···················· |
| H | 7260 | hCoV-19/England/QEUH-9A4032/2020\|EPI_ISL_540219\|20 | ···················· | ···················· | ···················· | ···················· | ···················· | ···················· |

| | | | | | | | | |
|---|---|---|---|---|---|---|---|---|
| H | 7261 hCoV-19/England/QEUH-9A458E/2020\|EPI_ISL_540220\|20 | ···················· | ···················· | ···················· | ···················· | ···················· | ···················· | |
| H | 7262 hCoV-19/England/QEUH-9A45E8/2020\|EPI_ISL_540221\|20 | ···················· | ···················· | ···················· | ···················· | ···T················ | ···················· | |
| H | 7263 hCoV-19/England/QEUH-9A41A8/2020\|EPI_ISL_540222\|20 | ···················· | ·A·················· | ···················· | ···················· | ···················· | ···················· | |
| H | 7264 hCoV-19/England/QEUH-9A42D2/2020\|EPI_ISL_540223\|20 | ···················· | ·A·················· | ···················· | ···················· | ···················· | ··A················· | |
| H | 7265 hCoV-19/England/QEUH-9A4612/2020\|EPI_ISL_540224\|20 | ···················· | ···················· | ···················· | ···················· | ···················· | ···················· | |
| H | 7266 hCoV-19/England/QEUH-9A42A5/2020\|EPI_ISL_540225\|20 | ···················· | ···················· | ···················· | ···················· | ···················· | ···················· | |
| H | 7267 hCoV-19/England/QEUH-9A422D/2020\|EPI_ISL_540226\|20 | ···················· | ···················· | ···················· | ···················· | ···················· | ···················· | |
| H | 7268 hCoV-19/England/QEUH-9A46C7/2020\|EPI_ISL_540227\|20 | ···················· | ···················· | ···················· | ···················· | ···················· | ···················· | |
| H | 7269 hCoV-19/England/QEUH-9A4551/2020\|EPI_ISL_540228\|20 | ···················· | ·A·················· | ···················· | ···················· | ···················· | ···················· | |
| H | 7270 hCoV-19/England/QEUH-9A407E/2020\|EPI_ISL_540230\|20 | ···················· | ···················· | ···················· | ···················· | ···················· | ···················· | |
| H | 7271 hCoV-19/England/QEUH-9A44EB/2020\|EPI_ISL_540231\|20 | ···················· | ···················· | ···················· | ···················· | ···················· | ···················· | |
| H | 7272 hCoV-19/Scotland/QEUH-96F73E/2020\|EPI_ISL_540233\|2 | ···················· | ···················· | ···················· | ···················· | ···················· | ···················· | |
| H | 7273 hCoV-19/England/QEUH-96F6E6/2020\|EPI_ISL_540234\|20 | ···················· | ···················· | ···················· | ···················· | ···················· | ···················· | |
| H | 7274 hCoV-19/England/QEUH-96F6F5/2020\|EPI_ISL_540235\|20 | ···················· | ···················· | ···················· | ···················· | ···················· | ···················· | |
| H | 7275 hCoV-19/Northern Ireland/QEUH-96FFEB/2020\|EPI_ISL_ | ···················· | ···················· | ···················· | ···················· | ···················· | ···················· | |
| H | 7276 hCoV-19/England/QEUH-96FADB/2020\|EPI_ISL_540239\|20 | ···················· | ···················· | ···················· | ···················· | ···················· | ···················· | |
| H | 7277 hCoV-19/Scotland/QEUH-96FDD2/2020\|EPI_ISL_540240\|2 | ···················· | ···················· | ···················· | ···················· | ···················· | ···················· | |
| H | 7278 hCoV-19/England/QEUH-9700B7/2020\|EPI_ISL_540242\|20 | ···················· | ···················· | ···················· | ···················· | ···················· | ···················· | |
| H | 7279 hCoV-19/Scotland/QEUH-96F798/2020\|EPI_ISL_540243\|2 | ···················· | ···················· | ···················· | ···················· | ···················· | ···················· | |
| H | 7280 hCoV-19/England/QEUH-96F7F2/2020\|EPI_ISL_540244\|20 | ···················· | ···················· | ···················· | ···················· | ···················· | ···················· | |
| H | 7281 hCoV-19/England/QEUH-96F631/2020\|EPI_ISL_540245\|20 | ···················· | ···················· | ···················· | ···················· | ···················· | ···················· | |
| H | 7282 hCoV-19/England/QEUH-96FA26/2020\|EPI_ISL_540246\|20 | ···················· | ···················· | ···················· | ···················· | ···················· | ···················· | |
| H | 7283 hCoV-19/England/QEUH-96F947/2020\|EPI_ISL_540248\|20 | ···················· | ···················· | ···················· | ···················· | ···················· | ···················· | |
| H | 7284 hCoV-19/Scotland/QEUH-96FC6C/2020\|EPI_ISL_540249\|2 | ···················· | ···················· | ···················· | ···················· | ···················· | ···················· | |
| H | 7285 hCoV-19/Northern Ireland/QEUH-96FF27/2020\|EPI_ISL_ | ···················· | ···················· | ···················· | ···················· | ···················· | ···················· | |
| H | 7286 hCoV-19/England/QEUH-96FA17/2020\|EPI_ISL_540252\|20 | ···················· | ···················· | ···················· | ···················· | ···················· | ···················· | |
| H | 7287 hCoV-19/England/QEUH-96FCC6/2020\|EPI_ISL_540253\|20 | ···················· | ···················· | ···················· | ···················· | ···················· | ···················· | |
| H | 7288 hCoV-19/Northern Ireland/QEUH-96FFFA/2020\|EPI_ISL_ | ···················· | ···················· | ···················· | ···················· | ···················· | ···················· | |
| H | 7289 hCoV-19/Scotland/QEUH-96F929/2020\|EPI_ISL_540256\|2 | ···················· | ···················· | ···················· | ···················· | ···················· | ···················· | |
| H | 7290 hCoV-19/Scotland/QEUH-96F604/2020\|EPI_ISL_540257\|2 | ···················· | ···················· | ···················· | ···················· | ···················· | ···················· | |
| H | 7291 hCoV-19/England/QEUH-96F7D4/2020\|EPI_ISL_540258\|20 | ···················· | ···················· | ···················· | ···················· | ···················· | ···················· | |
| H | 7292 hCoV-19/England/QEUH-96F868/2020\|EPI_ISL_540259\|20 | ···················· | ···················· | ···················· | ···················· | ···················· | ···················· | |
| H | 7293 hCoV-19/England/QEUH-97014B/2020\|EPI_ISL_540260\|20 | ···················· | ···················· | ···················· | ···················· | ···················· | ···················· | |
| H | 7294 hCoV-19/England/QEUH-9700D5/2020\|EPI_ISL_540261\|20 | ···················· | ···················· | ···················· | ···················· | ···················· | ···················· | |
| H | 7295 hCoV-19/England/QEUH-96FB32/2020\|EPI_ISL_540262\|20 | ···················· | ·A·················· | ···················· | ···················· | ···················· | ··A················· | |
| H | 7296 hCoV-19/England/QEUH-970169/2020\|EPI_ISL_540263\|20 | ···················· | ···················· | ···················· | ···················· | ···················· | ···················· | |
| H | 7297 hCoV-19/Scotland/QEUH-96FE75/2020\|EPI_ISL_540264\|2 | ···················· | ···················· | ···················· | ···················· | ···················· | ···················· | |
| H | 7298 hCoV-19/England/QEUH-96FACC/2020\|EPI_ISL_540265\|20 | ···················· | ···················· | ···················· | ···················· | ···················· | ···················· | |
| H | 7299 hCoV-19/England/QEUH-96F956/2020\|EPI_ISL_540266\|20 | ···················· | ·A·················· | ···················· | ···················· | ···················· | ···················· | |
| H | 7300 hCoV-19/Northern Ireland/QEUH-96FF63/2020\|EPI_ISL_ | ···················· | ···················· | ···················· | ···················· | ···················· | ···················· | |
| H | 7301 hCoV-19/England/QEUH-96F859/2020\|EPI_ISL_540268\|20 | ···················· | ·A·················· | ···················· | ···················· | ···················· | ···················· | |
| H | 7302 hCoV-19/Northern Ireland/QEUH-96FCB7/2020\|EPI_ISL_ | ···················· | ···················· | ···················· | ···················· | ···················· | ···················· | |
| H | 7303 hCoV-19/England/QEUH-96F77A/2020\|EPI_ISL_540270\|20 | ···················· | ···················· | ···················· | ···················· | ···················· | ···················· | |
| H | 7304 hCoV-19/Northern Ireland/QEUH-970002/2020\|EPI_ISL_ | ···················· | ···················· | ···················· | ···················· | ···················· | ···················· | |
| H | 7305 hCoV-19/England/QEUH-97011E/2020\|EPI_ISL_540272\|20 | ···················· | ···················· | ···················· | ···················· | ···················· | ···················· | |
| H | 7306 hCoV-19/England/QEUH-96F622/2020\|EPI_ISL_540274\|20 | ···················· | ···················· | ···················· | ···················· | ···················· | ···················· | |
| H | 7307 hCoV-19/England/QEUH-96FAF9/2020\|EPI_ISL_540276\|20 | ···················· | ···················· | ···················· | ···················· | ···················· | ···················· | |
| H | 7308 hCoV-19/Northern Ireland/QEUH-96FBAB/2020\|EPI_ISL_ | ···················· | ···················· | ···················· | ···················· | ···················· | ···················· | |
| H | 7309 hCoV-19/Northern Ireland/QEUH-96FB9C/2020\|EPI_ISL_ | ···················· | ···················· | ···················· | ···················· | ···················· | ···················· | |
| H | 7310 hCoV-19/Northern Ireland/QEUH-96FFCD/2020\|EPI_ISL_ | ···················· | ···················· | ···················· | ···················· | ···················· | ···················· | |
| H | 7311 hCoV-19/England/QEUH-96F91A/2020\|EPI_ISL_540282\|20 | ···················· | ···················· | ···················· | ···················· | ··················C· | ···················· | 1 |
| H | 7312 hCoV-19/England/QEUH-96FC8A/2020\|EPI_ISL_540283\|20 | ···················· | ···················· | ···················· | ···················· | ···················· | ···················· | |
| H | 7313 hCoV-19/England/QEUH-96F8E0/2020\|EPI_ISL_540285\|20 | ···················· | ···················· | ···················· | ···················· | ···················· | ···················· | |
| H | 7314 hCoV-19/Northern Ireland/QEUH-96FCA8/2020\|EPI_ISL_ | ···················· | ···················· | ···················· | ···················· | ···················· | ···················· | |
| H | 7315 hCoV-19/England/QEUH-97015A/2020\|EPI_ISL_540288\|20 | ···················· | ···················· | ···················· | ···················· | ···················· | ···················· | |
| H | 7316 hCoV-19/Switzerland/VD-ETHZ-270202/2020\|EPI_ISL_54 | ···················· | ···················· | ···················· | ···················· | ···················· | ···················· | |
| H | 7317 hCoV-19/Switzerland/VS-ETHZ-270074/2020\|EPI_ISL_54 | ···················· | ···················· | ···················· | ···················· | ···················· | ···················· | |
| H | 7318 hCoV-19/Switzerland/VS-ETHZ-270176/2020\|EPI_ISL_54 | ···················· | ···················· | ···················· | ···················· | ···················· | ···················· | |
| H | 7319 hCoV-19/USA/IL-UW-1488/2020\|EPI_ISL_541543\|2020-09 | ···················· | ·A·················· | ···················· | ···················· | ···················· | ···················· | |
| H | 7320 hCoV-19/USA/IL-UW-1490/2020\|EPI_ISL_541544\|2020-09 | ···················· | ·A·················· | ···················· | ···················· | ···················· | ···················· | |
| H | 7321 hCoV-19/USA/IL-UW-1494/2020\|EPI_ISL_541545\|2020-09 | ···················· | ···················· | ···················· | ···················· | ···················· | ···················· | |
| H | 7322 hCoV-19/USA/IL-UW-1503/2020\|EPI_ISL_541546\|2020-09 | ···················· | ···················· | ···················· | ···················· | ···················· | ···················· | |
| H | 7324 hCoV-19/USA/IL-UW-1521/2020\|EPI_ISL_541548\|2020-09 | ···················· | ···················· | ···················· | ···················· | ···················· | ···················· | |
| H | 7325 hCoV-19/USA/IL-UW-1522/2020\|EPI_ISL_541549\|2020-09 | ···················· | ···················· | ···················· | ···················· | ···················· | ···················· | |
| H | 7326 hCoV-19/USA/IL-UW-1524/2020\|EPI_ISL_541550\|2020-09 | ···················· | ···················· | ···················· | ···················· | ···················· | ···················· | |
| H | 7327 hCoV-19/USA/WI-UW-1418/2020\|EPI_ISL_541551\|2020-09 | ···················· | ···················· | ···················· | ···················· | ···················· | ···················· | |
| H | 7330 hCoV-19/USA/WI-UW-1421/2020\|EPI_ISL_541554\|2020-09 | ···················· | ···················· | ···················· | ···················· | ···················· | ···················· | |
| H | 7331 hCoV-19/USA/WI-UW-1422/2020\|EPI_ISL_541555\|2020-09 | ···················· | ·············A······ | ···················· | ···················· | ···················· | ···················· | |
| H | 7335 hCoV-19/USA/WI-UW-1426/2020\|EPI_ISL_541559\|2020-09 | ···················· | ···················· | ···················· | ···················· | ···················· | ···················· | |
| H | 7336 hCoV-19/USA/WI-UW-1427/2020\|EPI_ISL_541560\|2020-09 | ···················· | ···················· | ···················· | ···················· | ···················· | ···················· | |
| H | 7338 hCoV-19/USA/WI-UW-1429/2020\|EPI_ISL_541562\|2020-09 | ···················· | ···················· | ···················· | ···················· | ···················· | ···················· | |
| H | 7340 hCoV-19/USA/WI-UW-1431/2020\|EPI_ISL_541564\|2020-09 | ···················· | ···················· | ···················· | ···················· | ···················· | ···················· | |
| H | 7342 hCoV-19/USA/WI-UW-1433/2020\|EPI_ISL_541566\|2020-09 | ···················· | ···················· | ···················· | ···················· | ···················· | ···················· | |
| H | 7343 hCoV-19/USA/WI-UW-1434/2020\|EPI_ISL_541567\|2020-09 | ···················· | ···················· | ···················· | ···················· | ···················· | ···················· | |
| H | 7344 hCoV-19/USA/WI-UW-1436/2020\|EPI_ISL_541569\|2020-09 | ···················· | ···················· | ···················· | ···················· | ···················· | ···················· | |
| H | 7345 hCoV-19/USA/WI-UW-1437/2020\|EPI_ISL_541570\|2020-09 | ···················· | ···················· | ···················· | ···················· | ···················· | ···················· | |
| H | 7348 hCoV-19/USA/WI-UW-1442/2020\|EPI_ISL_541574\|2020-09 | ···················· | ···················· | ···················· | ···················· | ···················· | ···················· | |
| H | 7349 hCoV-19/USA/WI-UW-1443/2020\|EPI_ISL_541575\|2020-09 | ···················· | ···················· | ···················· | ···················· | ···················· | ···················· | |
| H | 7350 hCoV-19/USA/WI-UW-1444/2020\|EPI_ISL_541576\|2020-09 | ···················· | ···················· | ···················· | ···················· | ···················· | ···················· | |
| H | 7353 hCoV-19/USA/WI-UW-1447/2020\|EPI_ISL_541579\|2020-09 | ···················· | ···················· | ···················· | ···················· | ···················· | ···················· | |

| | | | | | | | | |
|---|---|---|---|---|---|---|---|---|
| H | 7354 | hCoV-19/USA/WI-UW-1448/2020\|EPI_ISL_541580\|2020-09 | ···················· | ···················· | ···················· | ···················· | ···················· | ···················· |
| H | 7355 | hCoV-19/USA/WI-UW-1449/2020\|EPI_ISL_541581\|2020-09 | ···················· | ·A·················· | ···················· | ···················· | ···················· | ···················· |
| H | 7356 | hCoV-19/USA/WI-UW-1450/2020\|EPI_ISL_541582\|2020-09 | ···················· | ···················· | ···················· | ···················· | ···················· | ···················· |
| H | 7357 | hCoV-19/USA/WI-UW-1451/2020\|EPI_ISL_541583\|2020-09 | ···················· | ···················· | ···················· | ···················· | ···················· | ···················· |
| H | 7359 | hCoV-19/USA/WI-UW-1453/2020\|EPI_ISL_541585\|2020-09 | ···················· | ···················· | ···················· | ···················· | ···················· | ···················· |
| H | 7360 | hCoV-19/USA/WI-UW-1454/2020\|EPI_ISL_541586\|2020-09 | ···················· | ···················· | ···················· | ···················· | ···················· | ···················· |
| H | 7361 | hCoV-19/USA/WI-UW-1455/2020\|EPI_ISL_541587\|2020-09 | ···················· | ···················· | ···················· | ···················· | ···················· | ···················· |
| H | 7362 | hCoV-19/USA/WI-UW-1456/2020\|EPI_ISL_541588\|2020-09 | ···················· | ···················· | ···················· | ···················· | ···················· | ···················· |
| H | 7364 | hCoV-19/USA/WI-UW-1458/2020\|EPI_ISL_541590\|2020-09 | ···················· | ·A·················· | ···················· | ···················· | ···················· | ···················· |
| H | 7366 | hCoV-19/USA/WI-UW-1460/2020\|EPI_ISL_541592\|2020-09 | ···················· | ···················· | ···················· | ···················· | ···················· | ···················· |
| H | 7367 | hCoV-19/USA/WI-UW-1461/2020\|EPI_ISL_541593\|2020-09 | ···················· | ···················· | ···················· | ···················· | ···················· | ···················· |
| H | 7368 | hCoV-19/USA/WI-UW-1462/2020\|EPI_ISL_541594\|2020-09 | ···················· | ···················· | ···················· | ···················· | ···················· | ···················· |
| H | 7369 | hCoV-19/USA/WI-UW-1463/2020\|EPI_ISL_541595\|2020-09 | ···················· | ···················· | ···················· | ···················· | ···················· | ···················· |
| H | 7372 | hCoV-19/USA/WI-UW-1466/2020\|EPI_ISL_541598\|2020-09 | ···················· | ···················· | ···················· | ···················· | ···················· | ···················· |
| H | 7373 | hCoV-19/USA/WI-UW-1467/2020\|EPI_ISL_541599\|2020-09 | ···················· | ···················· | ···················· | ···················· | ···················· | ···················· |
| H | 7374 | hCoV-19/USA/TX-HMH-1733/2020\|EPI_ISL_542800\|2020-0 | ···················· | ···················· | ···················· | ···················· | ···················· | ···················· |
| H | 7375 | hCoV-19/USA/TX-HMH-1734/2020\|EPI_ISL_542801\|2020-0 | ···················· | ···················· | ···················· | ···················· | ···················· | ···················· |
| H | 7376 | hCoV-19/USA/TX-HMH-1735/2020\|EPI_ISL_542802\|2020-0 | ···················· | ···················· | ···················· | ···················· | ···················· | ···················· |
| H | 7377 | hCoV-19/USA/TX-HMH-1736/2020\|EPI_ISL_542803\|2020-0 | ···················· | ···················· | ···················· | ···················· | ···················· | ···················· |
| H | 7379 | hCoV-19/USA/TX-HMH-1738/2020\|EPI_ISL_542805\|2020-0 | ···················· | ···················· | ···················· | ···················· | ···················· | ···················· |
| H | 7380 | hCoV-19/USA/TX-HMH-1739/2020\|EPI_ISL_542806\|2020-0 | ···················· | ···················· | ···················· | ···················· | ···················· | ···················· |
| H | 7381 | hCoV-19/USA/TX-HMH-1742/2020\|EPI_ISL_542808\|2020-0 | ···················· | ···················· | ···················· | ···················· | ···················· | ···················· |
| H | 7382 | hCoV-19/USA/TX-HMH-1744/2020\|EPI_ISL_542809\|2020-0 | ···················· | ···················· | ···················· | ···················· | ···················· | ···················· |
| H | 7383 | hCoV-19/USA/TX-HMH-1745/2020\|EPI_ISL_542810\|2020-0 | ···················· | ···················· | ···················· | ···················· | ···················· | ···················· |
| H | 7384 | hCoV-19/USA/TX-HMH-1746/2020\|EPI_ISL_542811\|2020-0 | ···················· | ···················· | ···················· | ···················· | ···················· | ···················· |
| H | 7385 | hCoV-19/USA/TX-HMH-1747/2020\|EPI_ISL_542812\|2020-0 | ···················· | ···················· | ···················· | ···················· | ···················· | ···················· |
| H | 7386 | hCoV-19/USA/TX-HMH-1750/2020\|EPI_ISL_542813\|2020-0 | ···················· | ···················· | ···················· | ···················· | ···················· | ···················· |
| H | 7387 | hCoV-19/USA/TX-HMH-1754/2020\|EPI_ISL_542814\|2020-0 | ···················· | ···················· | ···················· | ···················· | ···················· | ···················· |
| H | 7388 | hCoV-19/USA/TX-HMH-1755/2020\|EPI_ISL_542815\|2020-0 | ···················· | ···················· | ···················· | ···················· | ···················· | ···················· |
| H | 7389 | hCoV-19/USA/TX-HMH-1757/2020\|EPI_ISL_542817\|2020-0 | ···················· | ···················· | ···················· | ···················· | ···················· | ···················· |
| H | 7390 | hCoV-19/USA/TX-HMH-1758/2020\|EPI_ISL_542818\|2020-0 | ···················· | ···················· | ···················· | ···················· | ···················· | ···················· |
| H | 7392 | hCoV-19/USA/TX-HMH-1760/2020\|EPI_ISL_542820\|2020-0 | ···················· | ···················· | ···················· | ···················· | ···················· | ···················· |
| H | 7393 | hCoV-19/USA/TX-HMH-1761/2020\|EPI_ISL_542821\|2020-0 | ···················· | ···················· | ···················· | ···················· | ···················· | ···················· |
| H | 7394 | hCoV-19/USA/TX-HMH-1763/2020\|EPI_ISL_542822\|2020-0 | ···················· | ···················· | ···················· | ···················· | ···················· | ···················· |
| H | 7395 | hCoV-19/USA/TX-HMH-1764/2020\|EPI_ISL_542823\|2020-0 | ···················· | ···················· | ···················· | ···················· | ···················· | ···················· |
| H | 7396 | hCoV-19/USA/TX-HMH-1765/2020\|EPI_ISL_542824\|2020-0 | ···················· | ···················· | ···················· | ···················· | ··N················· | NN·················· |
| H | 7397 | hCoV-19/USA/TX-HMH-1766/2020\|EPI_ISL_542825\|2020-0 | ···················· | ···················· | ···················· | ···················· | ···················· | ···················· |
| H | 7398 | hCoV-19/USA/TX-HMH-1768/2020\|EPI_ISL_542826\|2020-0 | ···················· | ···················· | ···················· | ···················· | ···················· | ···················· |
| H | 7399 | hCoV-19/USA/TX-HMH-1769/2020\|EPI_ISL_542827\|2020-0 | ···················· | ···················· | ···················· | ···················· | ···················· | ···················· |
| H | 7400 | hCoV-19/USA/TX-HMH-1770/2020\|EPI_ISL_542828\|2020-0 | ···················· | ···················· | ···················· | ···················· | ···················· | ···A················ |
| H | 7401 | hCoV-19/USA/TX-HMH-1786/2020\|EPI_ISL_542838\|2020-0 | ···················· | ···················· | ···················· | ···················· | ···················· | ···················· |
| H | 7402 | hCoV-19/USA/TX-HMH-1788/2020\|EPI_ISL_542839\|2020-0 | ···················· | ···················· | ···················· | ···················· | ···················· | ···················· |
| H | 7403 | hCoV-19/USA/TX-HMH-1842/2020\|EPI_ISL_542875\|2020-0 | ···················· | ···················· | ···················· | ···················· | ···················· | ···················· |
| H | 7404 | hCoV-19/USA/TX-HMH-1844/2020\|EPI_ISL_542876\|2020-0 | ···················· | ···················· | ···················· | ···················· | ···················· | ···················· |
| H | 7405 | hCoV-19/USA/TX-HMH-1847/2020\|EPI_ISL_542878\|2020-0 | ···················· | ···················· | ···················· | ···················· | ···················· | ···················· |
| H | 7406 | hCoV-19/USA/TX-HMH-1853/2020\|EPI_ISL_542880\|2020-0 | ···················· | ···················· | ···················· | ···················· | ···················· | ···················· |
| H | 7407 | hCoV-19/USA/TX-HMH-1856/2020\|EPI_ISL_542881\|2020-0 | ···················· | ···················· | ···················· | ···················· | ···················· | ···················· |
| H | 7408 | hCoV-19/USA/TX-HMH-1860/2020\|EPI_ISL_542882\|2020-0 | ···················· | ···················· | ···················· | ···················· | ···················· | ···················· |
| H | 7409 | hCoV-19/USA/TX-HMH-1863/2020\|EPI_ISL_542883\|2020-0 | ···················· | ···················· | ···················· | ···················· | ···················· | ···················· |
| H | 7410 | hCoV-19/USA/TX-HMH-1865/2020\|EPI_ISL_542885\|2020-0 | ···················· | ···················· | ···················· | ···················· | ···················· | ···················· |
| H | 7411 | hCoV-19/USA/TX-HMH-1872/2020\|EPI_ISL_542886\|2020-0 | ···················· | ···················· | ···················· | ···················· | ···················· | ···················· |
| H | 7412 | hCoV-19/USA/TX-HMH-1873/2020\|EPI_ISL_542887\|2020-0 | ···················· | ···················· | ···················· | ···················· | ···················· | ···················· |
| H | 7413 | hCoV-19/USA/TX-HMH-1875/2020\|EPI_ISL_542888\|2020-0 | ···················· | ···················· | ···················· | ···················· | ···················· | ···················· |
| H | 7414 | hCoV-19/USA/TX-HMH-1877/2020\|EPI_ISL_542889\|2020-0 | ···················· | ··············C····· | ···················· | ···················· | ···················· | ···················· |
| H | 7415 | hCoV-19/USA/TX-HMH-1878/2020\|EPI_ISL_542890\|2020-0 | ···················· | ···················· | ···················· | ···················· | ···················· | ···················· |
| H | 7416 | hCoV-19/USA/TX-HMH-1880/2020\|EPI_ISL_542891\|2020-0 | ···················· | ···················· | ···················· | ···················· | ···················· | ···················· |
| H | 7417 | hCoV-19/USA/TX-HMH-1889/2020\|EPI_ISL_542892\|2020-0 | ···················· | ···················· | ···················· | ···················· | ···················· | ···················· |
| H | 7418 | hCoV-19/USA/TX-HMH-1895/2020\|EPI_ISL_542893\|2020-0 | ···················· | ···················· | ···················· | ···················· | ···················· | ···················· |
| H | 7419 | hCoV-19/USA/TX-HMH-1898/2020\|EPI_ISL_542894\|2020-0 | ···················· | ···················· | ···················· | ···················· | ···················· | ···················· |
| H | 7420 | hCoV-19/USA/TX-HMH-1899/2020\|EPI_ISL_542895\|2020-0 | ···················· | ······A············· | ···················· | ···················· | ···················· | ···················· |
| H | 7421 | hCoV-19/USA/TX-HMH-1903/2020\|EPI_ISL_542896\|2020-0 | ···················· | ···················· | ···················· | ···················· | ···················· | ···················· |
| H | 7422 | hCoV-19/USA/TX-HMH-1908/2020\|EPI_ISL_542897\|2020-0 | ···················· | ···················· | ···················· | ···················· | ···················· | ···················· |
| H | 7423 | hCoV-19/USA/TX-HMH-1910/2020\|EPI_ISL_542898\|2020-0 | ···················· | ···················· | ···················· | ···················· | ···················· | ···················· |
| H | 7424 | hCoV-19/USA/TX-HMH-1911/2020\|EPI_ISL_542899\|2020-0 | ···················· | ···················· | ···················· | ···················· | ···················· | ···················· |
| H | 7425 | hCoV-19/England/QEUH-96C3E2/2020\|EPI_ISL_530860\|20 | ···················· | ·A·················· | ···················· | ···················· | ···················· | ···················· |
| H | 7426 | hCoV-19/England/QEUH-96C652/2020\|EPI_ISL_530861\|20 | ···················· | ···················· | ···················· | ···················· | ···················· | ···················· |
| H | 7427 | hCoV-19/Scotland/QEUH-96C722/2020\|EPI_ISL_530862\|2 | ···················· | ···················· | ···················· | ···················· | ···················· | ···················· |
| H | 7428 | hCoV-19/England/QEUH-969C80/2020\|EPI_ISL_530863\|20 | ···················· | ·A·················· | ···················· | ···················· | ···················· | ···················· |
| H | 7429 | hCoV-19/England/QEUH-96C4A3/2020\|EPI_ISL_530864\|20 | ···················· | ···················· | ···················· | ···················· | ···················· | ···················· |
| H | 7430 | hCoV-19/England/QEUH-96C43A/2020\|EPI_ISL_530865\|20 | ···················· | ···················· | ···················· | ···················· | ···················· | ···················· |
| H | 7431 | hCoV-19/England/QEUH-96A0FC/2020\|EPI_ISL_530866\|20 | ···················· | ···················· | ···················· | ···················· | ···················· | ···················· |
| H | 7432 | hCoV-19/England/QEUH-96C1AC/2020\|EPI_ISL_530867\|20 | ···················· | ···················· | ···················· | ···················· | ···················· | ···················· |
| H | 7433 | hCoV-19/Scotland/QEUH-96C6BC/2020\|EPI_ISL_530869\|2 | ···················· | ···················· | ···················· | ···················· | ···················· | ···················· |
| H | 7434 | hCoV-19/Northern Ireland/QEUH-969D50/2020\|EPI_ISL_ | ···················· | ···················· | ···················· | ···················· | ···················· | ···················· |
| H | 7435 | hCoV-19/Scotland/QEUH-96C203/2020\|EPI_ISL_530871\|2 | ···················· | ···················· | ···················· | ···················· | ···················· | ···················· |
| H | 7436 | hCoV-19/England/QEUH-96C35B/2020\|EPI_ISL_530872\|20 | ···················· | ···················· | ···················· | ···················· | ···················· | ···················· |
| H | 7437 | hCoV-19/England/QEUH-96A0ED/2020\|EPI_ISL_530876\|20 | ···················· | ···················· | ···················· | ···················· | ···················· | ···················· |
| H | 7438 | hCoV-19/England/QEUH-96A1DB/2020\|EPI_ISL_530877\|20 | ···················· | ···················· | ···················· | ···················· | ···················· | ···················· |
| H | 7439 | hCoV-19/England/QEUH-96C40D/2020\|EPI_ISL_530878\|20 | ···················· | ···················· | ···················· | ···················· | ···················· | ···················· |
| H | 7440 | hCoV-19/England/QEUH-96C582/2020\|EPI_ISL_530879\|20 | ···················· | ···················· | ···················· | ···················· | ···················· | ···················· |

H 7441 hCoV-19/Northern Ireland/QEUH-969E02/2020|EPI_ISL_ ···················· ···················· ···················· ···················· ······T············· ····················
H 7442 hCoV-19/Northern Ireland/QEUH-96C009/2020|EPI_ISL_ ···················· ···················· ···················· ···················· ···················· ····················
H 7443 hCoV-19/England/QEUH-96C607/2020|EPI_ISL_530882|20 ···················· ·A·················· ···················· ···················· ···················· ··A·················
H 7444 hCoV-19/Northern Ireland/QEUH-96C045/2020|EPI_ISL_ ···················· ···················· ···················· ···················· ···················· ····················
H 7445 hCoV-19/Latvia/117/2020|EPI_ISL_534206|2020-08-05 ···················· ···················· ···················· ···················· ···················· ····················
H 7446 hCoV-19/Latvia/118/2020|EPI_ISL_534207|2020-07-29 ···················· ···················· ···················· ···················· ···················· ····················
H 7447 hCoV-19/Latvia/119/2020|EPI_ISL_534208|2020-07-29 ···················· ···················· ···················· ···················· ···················· ····················
H 7448 hCoV-19/Latvia/120/2020|EPI_ISL_534209|2020-07-31 ···················· ···················· ···················· ···················· ···················· ····················
H 7449 hCoV-19/Latvia/121/2020|EPI_ISL_534210|2020-08-24 ···················· ···················· ···················· ···················· ···················· ····················
H 7450 hCoV-19/Latvia/122/2020|EPI_ISL_534211|2020-08-24 ···················· ···················· ···················· ···················· ···················· ····················
H 7451 hCoV-19/Latvia/123/2020|EPI_ISL_534212|2020-08-22 ···················· ···················· ···················· ···················· ···················· ····················
H 7452 hCoV-19/Latvia/124/2020|EPI_ISL_534213|2020-08-24 ···················· ···················· ···················· ···················· ···················· ····················
H 7453 hCoV-19/Latvia/125/2020|EPI_ISL_534214|2020-08-25 ···················· ···················· ···················· ···················· ···················· ····················
H 7454 hCoV-19/Latvia/126/2020|EPI_ISL_534215|2020-08-25 ···················· ···················· ···················· ···················· ···················· ····················
H 7455 hCoV-19/Latvia/127/2020|EPI_ISL_534216|2020-08-25 ···················· ···················· ···················· ···················· ···················· ····················
H 7456 hCoV-19/Latvia/128/2020|EPI_ISL_534218|2020-08-25 ···················· ···················· ···················· ···················· ···················· ····················
H 7457 hCoV-19/Latvia/129/2020|EPI_ISL_534219|2020-08-25 ···················· ···················· ···················· ···················· ···················· ····················
H 7458 hCoV-19/Latvia/130/2020|EPI_ISL_534220|2020-08-25 ···················· ···················· ···················· ···················· ···················· ····················
H 7459 hCoV-19/Latvia/131/2020|EPI_ISL_534221|2020-08-24 ···················· ···················· ···················· ···················· ···················· ····················
H 7460 hCoV-19/Latvia/132/2020|EPI_ISL_534222|2020-08-22 ···················· ···················· ···················· ···················· ···················· ····················
H 7461 hCoV-19/Latvia/133/2020|EPI_ISL_534223|2020-08-26 ···················· ···················· ···················· ···················· ···················· ····················
H 7462 hCoV-19/Sweden/20-08927/2020|EPI_ISL_534224|2020-0 ···················· ···················· ···················· ···················· ···················· ····················
H 7463 hCoV-19/Sweden/20-08933/2020|EPI_ISL_534225|2020-0 ···················· ···················· ···················· ···················· ···················· ····················
H 7464 hCoV-19/Sweden/20-08934/2020|EPI_ISL_534226|2020-0 ···················· ···················· ···················· ···················· ···················· ····················
H 7465 hCoV-19/Sweden/20-08935/2020|EPI_ISL_534227|2020-0 ···················· ···················· ···················· ···················· ···················· ····················
H 7466 hCoV-19/Sweden/20-08938/2020|EPI_ISL_534228|2020-0 ···················· ···················· ···················· ···················· ···················· ····················
H 7467 hCoV-19/Sweden/20-08939/2020|EPI_ISL_534229|2020-0 ···················· ···················· ···················· ···················· ···················· ····················
H 7468 hCoV-19/Sweden/20-08947/2020|EPI_ISL_534230|2020-0 ···········T········ ···················· ···················· ···················· ···················· ····················
H 7469 hCoV-19/Sweden/20-08948/2020|EPI_ISL_534231|2020-0 ···················· ···················· ···················· ···················· ···················· ····················
H 7470 hCoV-19/Sweden/20-08950/2020|EPI_ISL_534232|2020-0 ···················· ···················· ···················· ···················· ···················· ····················
H 7471 hCoV-19/Sweden/20-08951/2020|EPI_ISL_534233|2020-0 ···················· ···················· ···················· ···················· ···················· ····················
H 7472 hCoV-19/Sweden/20-52284/2020|EPI_ISL_534234|2020-0 ···················· ···················· ···················· ···················· ···················· ····················
H 7473 hCoV-19/Sweden/20-52288/2020|EPI_ISL_534235|2020-0 ···················· ···················· ···················· ···················· ···················· ····················
H 7474 hCoV-19/Sweden/20-52289/2020|EPI_ISL_534236|2020-0 ···················· ···················· ·········T·········· ···················· ···················· ····················
H 7475 hCoV-19/Sweden/20-52304/2020|EPI_ISL_534237|2020-0 ···················· ···················· ···················· ···················· ···················· ····················
H 7476 hCoV-19/Sweden/20-52306/2020|EPI_ISL_534238|2020-0 ···················· ···················· ···················· ···················· ···················· ····················
H 7477 hCoV-19/Sweden/20-52384/2020|EPI_ISL_534239|2020-0 ···················· ···················· ···················· ···················· ···················· ····················
H 7478 hCoV-19/Sweden/20-52385/2020|EPI_ISL_534240|2020-0 ···················· ···················· ···················· ···················· ···················· ····················
H 7479 hCoV-19/Sweden/20-52386/2020|EPI_ISL_534241|2020-0 ···················· ···················· ···················· ···················· ···················· ····················
H 7480 hCoV-19/Sweden/20-52387/2020|EPI_ISL_534242|2020-0 ···················· ···················· ···················· ···················· ···················· ····················
H 7481 hCoV-19/Sweden/20-52388/2020|EPI_ISL_534243|2020-0 ···················· ···················· ···················· ···················· ···················· ····················
H 7482 hCoV-19/Sweden/20-52418/2020|EPI_ISL_534244|2020-0 ···················· ···················· ···················· ···················· ···················· ····················
H 7483 hCoV-19/Sweden/20-52425/2020|EPI_ISL_534245|2020-0 ···················· ···················· ···················· ···················· ···················· ····················
H 7484 hCoV-19/Sweden/20-52455/2020|EPI_ISL_534246|2020-0 ···················· ···················· ···················· ···················· ···················· ····················
H 7485 hCoV-19/Sweden/20-52462/2020|EPI_ISL_534247|2020-0 ···················· ···················· ···················· ···················· ···················· ····················
H 7486 hCoV-19/Sweden/20-52476/2020|EPI_ISL_534248|2020-0 ···················· ···················· ···················· ···················· ···················· ····················
H 7487 hCoV-19/Sweden/20-52533/2020|EPI_ISL_534249|2020-0 ···················· ···················· ···················· ···················· ···················· ····················
H 7488 hCoV-19/Sweden/20-52537/2020|EPI_ISL_534250|2020-0 ···················· ···················· ···················· ···················· ···················· ····················
H 7489 hCoV-19/Sweden/20-52538/2020|EPI_ISL_534251|2020-0 ···················· ···················· ···················· ···················· ···················· ····················
H 7490 hCoV-19/Sweden/20-52565/2020|EPI_ISL_534252|2020-0 ···················· ···················· ···················· ···················· ···················· ····················
H 7491 hCoV-19/Sweden/20-52566/2020|EPI_ISL_534253|2020-0 ···················· ···················· ···················· ···················· ···················· ····················
H 7492 hCoV-19/Sweden/20-52567/2020|EPI_ISL_534254|2020-0 ···················· ···················· ···················· ···················· ···················· ····················
H 7493 hCoV-19/Sweden/20-52568/2020|EPI_ISL_534255|2020-0 ···················· ···················· ···················· ···················· ···················· ····················
H 7494 hCoV-19/Sweden/20-52588/2020|EPI_ISL_534256|2020-0 ···················· ···················· ···················· ···················· ···················· ····················
H 7495 hCoV-19/Sweden/20-52637/2020|EPI_ISL_534257|2020-0 ···················· ···················· ···················· ···················· ···················· ····················
H 7496 hCoV-19/Sweden/20-52638/2020|EPI_ISL_534258|2020-0 ···················· ···················· ···················· ···················· ···················· ····················
H 7497 hCoV-19/Sweden/20-52647/2020|EPI_ISL_534259|2020-0 ···················· ···················· ···················· ···················· ···················· ····················
H 7498 hCoV-19/South Africa/KRISP-K002671/2020|EPI_ISL_53 ···················· ···················· ···················· ···················· ···················· ····················
H 7499 hCoV-19/South Africa/KRISP-K002673/2020|EPI_ISL_53 ···················· ···················· ···················· ···················· ···················· ····················
H 7500 hCoV-19/South Africa/KRISP-K002675/2020|EPI_ISL_53 ···················· ···················· ···················· ···················· ···················· ····················
H 7501 hCoV-19/South Africa/KRISP-K002676/2020|EPI_ISL_53 ···················· ···················· ···················· ···················· ···················· ····················
H 7502 hCoV-19/South Africa/KRISP-K002678/2020|EPI_ISL_53 ···················· ···················· ···················· ···················· ···················· ····················
H 7503 hCoV-19/South Africa/KRISP-K002679/2020|EPI_ISL_53 ···················· ···················· ···················· ···················· ···················· ····················
H 7504 hCoV-19/South Africa/KRISP-K002680/2020|EPI_ISL_53 ···················· ···················· ···················· ···················· ···················· ····················
H 7505 hCoV-19/South Africa/KRISP-K002681/2020|EPI_ISL_53 ···················· ···················· ···················· ···················· ···················· ····················
H 7506 hCoV-19/South Africa/KRISP-K002682/2020|EPI_ISL_53 ···················· ···················· ···················· ···················· ···················· ····················
H 7507 hCoV-19/South Africa/KRISP-K002684/2020|EPI_ISL_53 ···················· ···················· ···················· ···················· ···················· ····················
H 7508 hCoV-19/South Africa/KRISP-K002688/2020|EPI_ISL_53 ···················· ···················· ···················· ···················· ···T················ ····················
H 7509 hCoV-19/South Africa/KRISP-K002690/2020|EPI_ISL_53 ···················· ···················· ···················· ···················· ···················· ····················
H 7510 hCoV-19/South Africa/KRISP-K002691/2020|EPI_ISL_53 ···················· ···················· ···················· ···················· ···················· ····················
H 7511 hCoV-19/South Africa/KRISP-K002692/2020|EPI_ISL_53 ···················· ···················· ···················· ···················· ···················· ····················
H 7512 hCoV-19/South Africa/KRISP-K002693/2020|EPI_ISL_53 ···················· ···················· ···················· ···················· ···················· ····················
H 7513 hCoV-19/South Africa/KRISP-K002694/2020|EPI_ISL_53 ···················· ···················· ···················· ···················· ···················· ····················
H 7514 hCoV-19/South Africa/KRISP-K002696/2020|EPI_ISL_53 ···················· ···················· ···················· ···················· ···················· ····················
H 7515 hCoV-19/South Africa/KRISP-K002697/2020|EPI_ISL_53 ···················· ···················· ···················· ···················· ···················· ····················
H 7516 hCoV-19/South Africa/KRISP-K002698/2020|EPI_ISL_53 ···················· ···················· ···················· ···················· ···················· ····················
H 7517 hCoV-19/South Africa/KRISP-K002699/2020|EPI_ISL_53 ···················· ···················· ···················· ···················· ···················· ····················
H 7518 hCoV-19/South Africa/KRISP-K002700/2020|EPI_ISL_53 ···················· ···················· ···················· ···················· ···················· ····················
H 7519 hCoV-19/South Africa/KRISP-K002702/2020|EPI_ISL_53 ···················· ···················· ···················· ···················· ···················· ····················
H 7520 hCoV-19/South Africa/KRISP-K002703/2020|EPI_ISL_53 ···················· ···················· ···················· ···················· ···················· ····················

| | | | | | | | | | |
|---|---|---|---|---|---|---|---|---|---|
| H | 7521 | hCoV-19/South Africa/KRISP-K002704/2020\|EPI_ISL_53 | ···················· | ···················· | ····················· | ···················· | ···················· | ···················· | |
| H | 7522 | hCoV-19/South Africa/KRISP-K002706/2020\|EPI_ISL_53 | ···················· | ···················· | ····················· | ···················· | ···················· | ···················· | |
| H | 7523 | hCoV-19/South Africa/KRISP-K002707/2020\|EPI_ISL_53 | ···················· | ···················· | ····················· | ···················· | ···················· | ···················· | |
| H | 7524 | hCoV-19/South Africa/KRISP-K002708/2020\|EPI_ISL_53 | ···················· | ···················· | ····················· | ···················· | ···················· | ···················· | |
| H | 7525 | hCoV-19/South Africa/KRISP-K002709/2020\|EPI_ISL_53 | ···················· | ···················· | ····················· | ···················· | ···················· | ···················· | |
| H | 7526 | hCoV-19/South Africa/KRISP-K002710/2020\|EPI_ISL_53 | ···················· | ···················· | ····················· | ···················· | ···················· | ···················· | |
| H | 7527 | hCoV-19/South Africa/KRISP-K002711/2020\|EPI_ISL_53 | ···················· | ···················· | ····················· | ···················· | ···················· | ···················· | |
| H | 7528 | hCoV-19/South Africa/KRISP-K002712/2020\|EPI_ISL_53 | ···················· | ···················· | ····················· | ···················· | ···················· | ···················· | |
| H | 7529 | hCoV-19/South Africa/KRISP-K002715/2020\|EPI_ISL_53 | ···················· | ···················· | ····················· | ···················· | ···················· | ···················· | |
| H | 7530 | hCoV-19/Spain/CT-IrsiCaixa-2020090134/2020\|EPI_ISL | ···················· | ···················· | ····················· | ···················· | ···················· | ···················· | |
| H | 7531 | hCoV-19/Switzerland/BS-ETHZ-250030/2020\|EPI_ISL_53 | ···················· | ···················· | ····················· | ···················· | ···················· | ···················· | |
| H | 7532 | hCoV-19/Switzerland/AG-ETHZ-250060/2020\|EPI_ISL_53 | ···················· | ···················· | ····················· | ···················· | ···················· | ···················· | |
| H | 7533 | hCoV-19/Switzerland/BS-ETHZ-250061/2020\|EPI_ISL_53 | ···················· | ···················· | ····················· | ···················· | ···················· | ···················· | |
| H | 7534 | hCoV-19/Switzerland/BE-ETHZ-250062/2020\|EPI_ISL_53 | ···················· | ···················· | ····················· | ···················· | ···················· | ···················· | |
| H | 7535 | hCoV-19/Switzerland/BE-ETHZ-250063/2020\|EPI_ISL_53 | ···················· | ···················· | ····················· | ···················· | ···················· | ···················· | |
| H | 7536 | hCoV-19/Switzerland/BL-ETHZ-250064/2020\|EPI_ISL_53 | ···················· | ·A·················· | ····················· | ···················· | ···················· | ···················· | |
| H | 7537 | hCoV-19/Switzerland/GR-ETHZ-250068/2020\|EPI_ISL_53 | ···················· | ···················· | ····················· | ···················· | ···················· | ···················· | |
| H | 7538 | hCoV-19/Switzerland/ZH-ETHZ-250070/2020\|EPI_ISL_53 | ···················· | ···················· | ····················· | ···················· | ···················· | ···················· | |
| H | 7539 | hCoV-19/Switzerland/VD-ETHZ-250071/2020\|EPI_ISL_53 | ···················· | ···················· | ····················· | ···················· | ···················· | ···················· | |
| H | 7540 | hCoV-19/Switzerland/UR-ETHZ-250072/2020\|EPI_ISL_53 | ···················· | ···················· | ····················· | ···················· | ···················· | ···················· | |
| H | 7541 | hCoV-19/Switzerland/BL-ETHZ-250075/2020\|EPI_ISL_53 | ···················· | ···················· | ····················· | ···················· | ···················· | ···················· | |
| H | 7542 | hCoV-19/Switzerland/BL-ETHZ-250076/2020\|EPI_ISL_53 | ···················· | ···················· | ····················· | ···················· | ···················· | ···················· | |
| H | 7543 | hCoV-19/Northern Ireland/QEUH-9650D3/2020\|EPI_ISL_ | ···················· | ···················· | ····················· | ···················· | ···················· | ···················· | |
| H | 7544 | hCoV-19/England/QEUH-9640A7/2020\|EPI_ISL_536869\|20 | ···················· | ···················· | ····················· | ···················· | ···················· | ···················· | |
| H | 7545 | hCoV-19/Northern Ireland/QEUH-96510D/2020\|EPI_ISL_ | ···················· | ···················· | ····················· | ···················· | ···················· | ···················· | |
| H | 7546 | hCoV-19/Northern Ireland/QEUH-96406B/2020\|EPI_ISL_ | ···················· | ···················· | ····················· | ···················· | ···················· | ···················· | |
| H | 7547 | hCoV-19/England/QEUH-96A2E7/2020\|EPI_ISL_536872\|20 | ···················· | ···················· | ····················· | ···················· | ···················· | ···················· | |
| H | 7548 | hCoV-19/Northern Ireland/QEUH-963F05/2020\|EPI_ISL_ | ···················· | ···················· | ····················· | ···················· | ···················· | ···················· | |
| H | 7549 | hCoV-19/Northern Ireland/QEUH-964502/2020\|EPI_ISL_ | ···················· | ···················· | ····················· | ···················· | ···················· | ···················· | |
| H | 7550 | hCoV-19/England/QEUH-96463C/2020\|EPI_ISL_536875\|20 | ···················· | ···················· | ····················· | ···················· | ···················· | ···················· | |
| H | 7551 | hCoV-19/England/QEUH-9643EA/2020\|EPI_ISL_536876\|20 | ···················· | ···················· | ····················· | ···················· | ···················· | ···················· | |
| H | 7552 | hCoV-19/England/QEUH-9642DE/2020\|EPI_ISL_536877\|20 | ···················· | ···················· | ····················· | ···················· | ···················· | ···················· | |
| H | 7553 | hCoV-19/England/QEUH-9645C6/2020\|EPI_ISL_536879\|20 | ···················· | ···················· | ····················· | ···················· | ···················· | ···················· | |
| H | 7554 | hCoV-19/England/QEUH-964423/2020\|EPI_ISL_536880\|20 | ···················· | ···················· | ····················· | ···················· | ···················· | ···················· | |
| H | 7555 | hCoV-19/England/QEUH-9643AE/2020\|EPI_ISL_536881\|20 | ···················· | ···················· | ····················· | ···················· | ···················· | ···················· | |
| H | 7556 | hCoV-19/England/QEUH-964678/2020\|EPI_ISL_536882\|20 | ···················· | ···················· | ····················· | ···················· | ···················· | ···················· | |
| H | 7557 | hCoV-19/Northern Ireland/QEUH-9640B6/2020\|EPI_ISL_ | ···················· | ···················· | ····················· | ···················· | ···················· | ···················· | |
| H | 7558 | hCoV-19/England/QEUH-96446F/2020\|EPI_ISL_536885\|20 | ···················· | ···················· | ····················· | ···················· | ···················· | ···················· | |
| H | 7559 | hCoV-19/Northern Ireland/QEUH-963FE7/2020\|EPI_ISL_ | ···················· | ···················· | ····················· | ···················· | ···················· | ···················· | |
| H | 7560 | hCoV-19/England/QEUH-96421A/2020\|EPI_ISL_536887\|20 | ···················· | ···················· | ····················· | ···················· | ···················· | ···················· | |
| H | 7561 | hCoV-19/England/QEUH-964177/2020\|EPI_ISL_536888\|20 | ···················· | ···················· | ····················· | ···················· | ···················· | ···················· | |
| H | 7562 | hCoV-19/Northern Ireland/QEUH-963EDB/2020\|EPI_ISL_ | ···················· | ···················· | ····················· | ···················· | ···················· | ···················· | |
| H | 7563 | hCoV-19/England/QEUH-964247/2020\|EPI_ISL_536891\|20 | ···················· | ···················· | ····················· | ···················· | ···················· | ···················· | |
| H | 7564 | hCoV-19/England/QEUH-9642ED/2020\|EPI_ISL_536892\|20 | ···················· | ···················· | ····················· | ···················· | ···················· | ···················· | |
| H | 7565 | hCoV-19/Northern Ireland/QEUH-965158/2020\|EPI_ISL_ | ···················· | ···················· | ····················· | ···················· | ···················· | ···················· | |
| H | 7566 | hCoV-19/England/QEUH-964818/2020\|EPI_ISL_536894\|20 | ···················· | ···················· | ····················· | ···················· | ···················· | ···················· | |
| H | 7567 | hCoV-19/Northern Ireland/QEUH-963EEA/2020\|EPI_ISL_ | ···················· | ···················· | ····················· | ···················· | ···················· | ···················· | |
| H | 7568 | hCoV-19/England/QEUH-964696/2020\|EPI_ISL_536896\|20 | ···················· | ···················· | ····················· | ···················· | ···················· | ···················· | |
| H | 7569 | hCoV-19/England/QEUH-9643DB/2020\|EPI_ISL_536897\|20 | ···················· | ···················· | ····················· | ···················· | ···················· | ···················· | |
| H | 7570 | hCoV-19/Northern Ireland/QEUH-965149/2020\|EPI_ISL_ | ···················· | ···················· | ····················· | ···················· | ···················· | ···················· | |
| H | 7571 | hCoV-19/England/QEUH-964229/2020\|EPI_ISL_536899\|20 | ···················· | ···················· | ····················· | ···················· | ···················· | ···················· | |
| H | 7572 | hCoV-19/England/LEED-2A91E6/2020\|EPI_ISL_539061\|20 | ···················· | ···················· | ····················· | ···················· | ···················· | ···················· | |
| H | 7573 | hCoV-19/England/LEED-2A8132/2020\|EPI_ISL_539063\|20 | ···················· | ···················· | ····················· | ···················· | ···················· | ···················· | |
| H | 7574 | hCoV-19/England/LEED-2A8590/2020\|EPI_ISL_539064\|20 | ···················· | ···················· | ····················· | ···················· | ···················· | ···················· | |
| H | 7575 | hCoV-19/England/LEED-2A8BBC/2020\|EPI_ISL_539065\|20 | ···················· | ···················· | ····················· | ···················· | ···················· | ···················· | |
| H | 7576 | hCoV-19/England/LEED-2A835A/2020\|EPI_ISL_539067\|20 | ···················· | ···················A | ····················· | ···················· | ···················· | ···················· | 1 |
| H | 7577 | hCoV-19/England/LEED-2A8C7D/2020\|EPI_ISL_539068\|20 | ···················· | ···················· | ····················· | ···················· | ···················· | ···················· | |
| H | 7578 | hCoV-19/England/LEED-2A82A8/2020\|EPI_ISL_539069\|20 | ···················· | ···················· | ····················· | ···················· | ···················· | ···················· | |
| H | 7579 | hCoV-19/England/LEED-2A8F56/2020\|EPI_ISL_539071\|20 | ···················· | ·A·················· | ····················· | ···················· | ···················· | ···················· | |
| H | 7580 | hCoV-19/England/LEED-2A8BE9/2020\|EPI_ISL_539072\|20 | ···················· | ···················· | ····················· | ···················· | ···················· | ···················· | |
| H | 7581 | hCoV-19/England/LEED-2A89B2/2020\|EPI_ISL_539073\|20 | ···················· | ···················· | ····················· | ···················· | ···················· | ···················· | |
| H | 7582 | hCoV-19/England/LEED-2A875E/2020\|EPI_ISL_539078\|20 | ···················· | ·············A······ | ····················· | ···················· | ···················· | ···················· | |
| H | 7583 | hCoV-19/England/LEED-2A9131/2020\|EPI_ISL_539079\|20 | ···················· | ···················· | ····················· | ···················· | ···················· | ···················· | |
| H | 7584 | hCoV-19/England/LEED-2A841B/2020\|EPI_ISL_539200\|20 | ···················· | ···················· | ····················· | ···················· | ···················· | ···················· | |
| H | 7585 | hCoV-19/England/LEED-2A934A/2020\|EPI_ISL_539202\|20 | ···················· | ···················· | ····················· | ···················· | ···················· | ···················· | |
| H | 7586 | hCoV-19/England/LEED-2A84C0/2020\|EPI_ISL_539203\|20 | ···················· | ···················· | ····················· | ···················· | ···················· | ···················· | |
| H | 7587 | hCoV-19/England/LEED-2A9377/2020\|EPI_ISL_539205\|20 | ···················· | ···················· | ····················· | ···················· | ···················· | ···················· | |
| H | 7588 | hCoV-19/England/LEED-2A840C/2020\|EPI_ISL_539206\|20 | ···················· | ···················· | ····················· | ···················· | ···················· | ···················· | |
| H | 7589 | hCoV-19/England/LEED-2A9359/2020\|EPI_ISL_539207\|20 | ···················· | ···················· | ····················· | ···················· | ···················· | ···················· | |
| H | 7590 | hCoV-19/England/LEED-2A8572/2020\|EPI_ISL_539208\|20 | ···················· | ···················· | ····················· | ···················· | ···················· | ···················· | |
| H | 7591 | hCoV-19/England/LEED-2A92F2/2020\|EPI_ISL_539209\|20 | ···················· | ···················· | ····················· | ···················· | ···················· | ···················· | |
| H | 7592 | hCoV-19/England/LEED-2A8E3B/2020\|EPI_ISL_539211\|20 | ···················· | ···················· | ····················· | ···················· | ···················· | ···················· | |
| H | 7593 | hCoV-19/England/LEED-2A926B/2020\|EPI_ISL_539212\|20 | ···················· | ···················A | ····················· | ···················· | ···················· | ···················· | 1 |
| H | 7594 | hCoV-19/England/LEED-2A924D/2020\|EPI_ISL_539214\|20 | ···················· | ···················· | ····················· | ···················· | ···················· | ···················· | |
| H | 7595 | hCoV-19/England/LEED-2A8439/2020\|EPI_ISL_539215\|20 | ···················· | ···················· | ····················· | ···················· | ···················· | ···················· | |
| H | 7596 | hCoV-19/England/LEED-2A930E/2020\|EPI_ISL_539216\|20 | ···················· | ···················· | ····················· | ···················· | ···················· | ···················· | |
| H | 7597 | hCoV-19/England/LEED-2A8E2C/2020\|EPI_ISL_539217\|20 | ···················· | ···················· | ····················· | ···················· | ···················· | ···················· | |
| H | 7598 | hCoV-19/England/LEED-2A8DF2/2020\|EPI_ISL_539218\|20 | ···················· | ···················· | ····················· | ···················· | ···················· | ···················· | |
| H | 7599 | hCoV-19/England/LEED-2A874F/2020\|EPI_ISL_539219\|20 | ···················· | ···················· | ····················· | ···················· | ···················· | ···················· | |
| H | 7600 | hCoV-19/England/QEUH-98FC93/2020\|EPI_ISL_540047\|20 | ···················· | ·A·················· | ····················· | ···················· | ···················· | ···A················ | |

| | | | | | | | |
|---|---|---|---|---|---|---|---|
| H | 7601 hCoV-19/England/QEUH-9986C6/2020\|EPI_ISL_540048\|20 | ···················· | ···················· | ····················· | ···················· | ···················· | ···················· |
| H | 7602 hCoV-19/Northern Ireland/QEUH-98FC66/2020\|EPI_ISL_ | ···················· | ···················· | ····················· | ···················· | ···················· | ···················· |
| H | 7603 hCoV-19/England/QEUH-998E55/2020\|EPI_ISL_540051\|20 | ···················· | ···················· | ····················· | ···················· | ···················· | ···················· |
| H | 7604 hCoV-19/England/QEUH-990400/2020\|EPI_ISL_540052\|20 | ···················· | ·A·················· | ····················· | ···················· | ···················· | ··A················· |
| H | 7605 hCoV-19/England/QEUH-9901FA/2020\|EPI_ISL_540054\|20 | ···················· | ···················· | ····················· | ···················· | ···················· | ···················· |
| H | 7606 hCoV-19/England/QEUH-998893/2020\|EPI_ISL_540055\|20 | ···················· | ···················· | ····················· | ···················· | ···················· | ···················· |
| H | 7607 hCoV-19/England/QEUH-998BF4/2020\|EPI_ISL_540056\|20 | ···················· | ·A·················· | ····················· | ···················· | ···················· | ···················· |
| H | 7608 hCoV-19/England/QEUH-998AE8/2020\|EPI_ISL_540057\|20 | ···················· | ···················· | ····················· | ···················· | ···················· | ···················· |
| H | 7609 hCoV-19/England/QEUH-98FFD6/2020\|EPI_ISL_540058\|20 | ···················· | ···················· | ····················· | ···················· | ···················· | ···················· |
| H | 7610 hCoV-19/England/QEUH-99870F/2020\|EPI_ISL_540065\|20 | ···················· | ···················· | ····················· | ···················· | ···················· | ···················· |
| H | 7611 hCoV-19/England/QEUH-98FEBB/2020\|EPI_ISL_540067\|20 | ···················· | ·A·················· | ····················· | ···················· | ···················· | ···················· |
| H | 7612 hCoV-19/England/QEUH-998C3D/2020\|EPI_ISL_540068\|20 | ···················· | ···················· | ····················· | ···················· | ···················· | ···················· |
| H | 7613 hCoV-19/England/QEUH-9A44DC/2020\|EPI_ISL_540200\|20 | ···················· | ···················· | ····················· | ···················· | ···················· | ···················· |
| H | 7614 hCoV-19/Brazil/SE-6544/2020\|EPI_ISL_541378\|2020-04 | ···················· | ···················· | ····················· | ···················· | ···················· | ···················· |
| H | 7615 hCoV-19/Brazil/SE-6549/2020\|EPI_ISL_541379\|2020-04 | ···················· | ···················· | ····················· | ···················· | ···················· | ···················· |
| H | 7616 hCoV-19/Brazil/SE-6550/2020\|EPI_ISL_541380\|2020-04 | ···················· | ···················· | ····················· | ···················· | ···················· | ···················· |
| H | 7617 hCoV-19/Brazil/SE-6555/2020\|EPI_ISL_541381\|2020-04 | ···················· | ···················· | ····················· | ···················· | ···················· | ···················· |
| H | 7618 hCoV-19/Brazil/SE-6556/2020\|EPI_ISL_541382\|2020-04 | ···················· | ···················· | ····················· | ···················· | ···················· | ···················· |
| H | 7619 hCoV-19/Brazil/SE-6557/2020\|EPI_ISL_541383\|2020-04 | ···················· | ···················· | ····················· | ···················· | ···················· | ···················· |
| H | 7620 hCoV-19/Brazil/SE-6561/2020\|EPI_ISL_541384\|2020-04 | ···················· | ···················· | ····················· | ···················· | ···················· | ···················· |
| H | 7621 hCoV-19/Brazil/SE-6563/2020\|EPI_ISL_541385\|2020-04 | ···················· | ···················· | ····················· | ···················· | ···················· | ···················· |
| H | 7622 hCoV-19/Brazil/SE-6567/2020\|EPI_ISL_541386\|2020-04 | ···················· | ···················· | ····················· | ···················· | ···················· | ···················· |
| H | 7623 hCoV-19/Brazil/SE-6568/2020\|EPI_ISL_541387\|2020-04 | ···················· | ···················· | ····················· | ···················· | ···················· | ···················· |
| H | 7624 hCoV-19/Brazil/SE-6579/2020\|EPI_ISL_541388\|2020-04 | ···················· | ···················· | ····················· | ···················· | ···················· | ···················· |
| H | 7625 hCoV-19/Brazil/SE-6583/2020\|EPI_ISL_541389\|2020-04 | ···················· | ···················· | ····················· | ···················· | ···················· | ···················· |
| H | 7626 hCoV-19/Brazil/SE-6608/2020\|EPI_ISL_541396\|2020-04 | ···················· | ···················· | ····················· | ···················· | ···················· | ···················· |
| H | 7627 hCoV-19/Switzerland/VD-ETHZ-270177/2020\|EPI_ISL_54 | ···················· | ···················· | ····················· | ···················· | ···················· | ···················· |
| H | 7628 hCoV-19/Switzerland/VD-ETHZ-270187/2020\|EPI_ISL_54 | ···················· | ···················· | ····················· | ···················· | ···················· | ···················· |
| H | 7629 hCoV-19/Switzerland/VD-ETHZ-270199/2020\|EPI_ISL_54 | ···················· | ···················· | ····················· | ···················· | ···················· | ···················· |
| H | 7630 hCoV-19/Switzerland/VD-ETHZ-270200/2020\|EPI_ISL_54 | ···················· | ···················· | ····················· | ···················· | ···················· | ···················· |
| H | 7631 hCoV-19/Switzerland/VD-ETHZ-270201/2020\|EPI_ISL_54 | ···················· | ···················· | ····················· | ···················· | ···················· | ···················· |
| H | 7632 hCoV-19/Switzerland/ZH-ETHZ-270039/2020\|EPI_ISL_54 | ···················· | ·A·················· | ····················· | ···················· | ···················· | ···················· |
| H | 7633 hCoV-19/Switzerland/ZH-ETHZ-270040/2020\|EPI_ISL_54 | ···················· | ···················· | ····················· | ···················· | ···················· | ···················· |
| H | 7634 hCoV-19/Switzerland/ZH-ETHZ-270044/2020\|EPI_ISL_54 | ···················· | ···················· | ····················· | ···················· | ···················· | ···················· |
| H | 7635 hCoV-19/Switzerland/ZH-ETHZ-270047/2020\|EPI_ISL_54 | ···················· | ···················· | ····················· | ···················· | ···················· | ···················· |
| H | 7636 hCoV-19/Switzerland/ZH-ETHZ-270051/2020\|EPI_ISL_54 | ···················· | ···················· | ····················· | ···················· | ···················· | ···················· |
| H | 7637 hCoV-19/Switzerland/ZH-ETHZ-270069/2020\|EPI_ISL_54 | ···················· | ···················· | ····················· | ···················· | ···················· | ···················· |
| H | 7638 hCoV-19/Switzerland/ZH-ETHZ-270081/2020\|EPI_ISL_54 | ···················· | ···················· | ····················· | ···················· | ···················· | ···················· |
| H | 7639 hCoV-19/Switzerland/ZH-ETHZ-270084/2020\|EPI_ISL_54 | ···················· | ···················· | ····················· | ···················· | ······K············· | ···················· |
| H | 7640 hCoV-19/Switzerland/ZH-ETHZ-270086/2020\|EPI_ISL_54 | ···················· | ···················· | ····················· | ···················· | ············T······· | ···················· |
| H | 7641 hCoV-19/Switzerland/ZH-ETHZ-270099/2020\|EPI_ISL_54 | ···················· | ···················· | ····················· | ···················· | ···················· | ···················· |
| H | 7642 hCoV-19/Switzerland/ZH-ETHZ-270100/2020\|EPI_ISL_54 | ···················· | ···················· | ····················· | ···················· | ···················· | ···················· |
| H | 7643 hCoV-19/Switzerland/ZH-ETHZ-270118/2020\|EPI_ISL_54 | ···················· | ···················· | ····················· | ···················· | ···················· | ···················· |
| H | 7644 hCoV-19/Switzerland/ZH-ETHZ-270119/2020\|EPI_ISL_54 | ···················· | ·A·················· | ····················· | ···················· | ···················· | ···················· |
| H | 7645 hCoV-19/Switzerland/ZH-ETHZ-270128/2020\|EPI_ISL_54 | ···················· | ·A·················· | ····················· | ···················· | ···················· | ············R······· |
| H | 7646 hCoV-19/Switzerland/ZH-ETHZ-270142/2020\|EPI_ISL_54 | ···················· | ···················· | ····················· | ···················· | ···················· | ···················· |
| H | 7647 hCoV-19/Switzerland/ZH-ETHZ-270143/2020\|EPI_ISL_54 | ···················· | ···················· | ····················· | ···················· | ···················· | ···················· |
| H | 7648 hCoV-19/Switzerland/ZH-ETHZ-270144/2020\|EPI_ISL_54 | ···················· | ···················· | ····················· | ···················· | ···················· | ···················· |
| H | 7649 hCoV-19/Switzerland/ZH-ETHZ-270146/2020\|EPI_ISL_54 | ···················· | ···················· | ····················· | ···················· | ···················· | ···················· |
| H | 7650 hCoV-19/Switzerland/ZH-ETHZ-270147/2020\|EPI_ISL_54 | ···················· | ···················· | ····················· | ···················· | ···················· | ···················· |
| H | 7651 hCoV-19/Switzerland/ZH-ETHZ-270170/2020\|EPI_ISL_54 | ···················· | ···················· | ····················· | ···················· | ···················· | ···················· |
| H | 7652 hCoV-19/Switzerland/ZH-ETHZ-270183/2020\|EPI_ISL_54 | ···················· | ···················· | ····················· | ···················· | ···················· | ···················· |
| H | 7653 hCoV-19/Switzerland/ZH-ETHZ-270184/2020\|EPI_ISL_54 | ···················· | ···················· | ····················· | ···················· | ···················· | ···················· |
| H | 7654 hCoV-19/USA/TX-HMH-1774/2020\|EPI_ISL_542830\|2020-0 | ···················· | ···················· | ····················· | ···················· | ···················· | ···················· |
| H | 7655 hCoV-19/USA/TX-HMH-1776/2020\|EPI_ISL_542831\|2020-0 | ···················· | ···················· | ····················· | ···················· | ···················· | ···················· |
| H | 7656 hCoV-19/USA/TX-HMH-1777/2020\|EPI_ISL_542832\|2020-0 | ···················· | ···················· | ····················· | ···················· | ···················· | ···················· |
| H | 7657 hCoV-19/USA/TX-HMH-1778/2020\|EPI_ISL_542833\|2020-0 | ···················· | ···················· | ····················· | ···················· | ···················· | ···················· |
| H | 7658 hCoV-19/USA/TX-HMH-1780/2020\|EPI_ISL_542834\|2020-0 | ···················· | ···················· | ····················· | ···················· | ···················· | ···················· |
| H | 7659 hCoV-19/USA/TX-HMH-1781/2020\|EPI_ISL_542835\|2020-0 | ···················· | ···················· | ····················· | ···················· | ···················· | ···················· |
| H | 7660 hCoV-19/USA/TX-HMH-1782/2020\|EPI_ISL_542836\|2020-0 | ···················· | ···················· | ····················· | ···················· | ···················· | ··A················· |
| H | 7661 hCoV-19/USA/TX-HMH-1789/2020\|EPI_ISL_542840\|2020-0 | ···················· | ···················· | ····················· | ···················· | ···················· | ···················· |
| H | 7662 hCoV-19/USA/TX-HMH-1790/2020\|EPI_ISL_542841\|2020-0 | ···················· | ···················· | ····················· | ···················· | ···················· | ···················· |
| H | 7664 hCoV-19/USA/TX-HMH-1794/2020\|EPI_ISL_542843\|2020-0 | ···················· | ···················· | ····················· | ···················· | ···················· | ···················· |
| H | 7665 hCoV-19/USA/TX-HMH-1795/2020\|EPI_ISL_542844\|2020-0 | ···················· | ···················· | ····················· | ···················· | ···················· | ···················· |
| H | 7666 hCoV-19/USA/TX-HMH-1796/2020\|EPI_ISL_542845\|2020-0 | ···················· | ···················· | ····················· | ···················· | ···················· | ···················· |
| H | 7667 hCoV-19/USA/TX-HMH-1797/2020\|EPI_ISL_542846\|2020-0 | ···················· | ···················· | ····················· | ···················· | ···················· | ···················· |
| H | 7668 hCoV-19/USA/TX-HMH-1798/2020\|EPI_ISL_542847\|2020-0 | ···················· | ···················· | ····················· | ···················· | ···················· | ···················· |
| H | 7669 hCoV-19/USA/TX-HMH-1799/2020\|EPI_ISL_542848\|2020-0 | ···················· | ···················· | ····················· | ···················· | ···················· | ···················· |
| H | 7672 hCoV-19/USA/TX-HMH-1803/2020\|EPI_ISL_542851\|2020-0 | ···················· | ···················· | ····················· | ···················· | ···················· | ···················· |
| H | 7673 hCoV-19/USA/TX-HMH-1804/2020\|EPI_ISL_542852\|2020-0 | ···················· | ···················· | ····················· | ···················· | ···················· | ···················· |
| H | 7674 hCoV-19/USA/TX-HMH-1806/2020\|EPI_ISL_542854\|2020-0 | ···················· | ···················· | ····················· | ···················· | ···················· | ···················· |
| H | 7675 hCoV-19/USA/TX-HMH-1807/2020\|EPI_ISL_542855\|2020-0 | ···················· | ···················· | ····················· | ···················· | ···················· | ···················· |
| H | 7676 hCoV-19/USA/TX-HMH-1812/2020\|EPI_ISL_542856\|2020-0 | ···················· | ···················· | ····················· | ···················· | ···················· | ···················· |
| H | 7677 hCoV-19/USA/TX-HMH-1813/2020\|EPI_ISL_542857\|2020-0 | ···················· | ···················· | ····················· | ···················· | ···················· | ···················· |
| H | 7678 hCoV-19/USA/TX-HMH-1815/2020\|EPI_ISL_542859\|2020-0 | ···················· | ···················· | ····················· | ···················· | ···················· | ···················· |
| H | 7679 hCoV-19/USA/TX-HMH-1817/2020\|EPI_ISL_542860\|2020-0 | ···················· | ··············C····· | ····················· | ···················· | ···················· | ···················· |
| H | 7680 hCoV-19/USA/TX-HMH-1819/2020\|EPI_ISL_542862\|2020-0 | ···················· | ···················· | ····················· | ···················· | ···················· | ···················· |
| H | 7681 hCoV-19/Northern Ireland/QEUH-96CB80/2020\|EPI_ISL_ | ···················· | ···················· | ····················· | ···················· | ···················· | ···················· |
| H | 7682 hCoV-19/Scotland/QEUH-96D378/2020\|EPI_ISL_530764\|2 | ···················· | ···················· | ····················· | ···················· | ···················· | ···················· |
| H | 7683 hCoV-19/Northern Ireland/QEUH-96D008/2020\|EPI_ISL_ | ···················· | ···················· | ····················· | ···················· | ···················· | ···················· |

| | | | | | | | | |
|---|---|---|---|---|---|---|---|---|
| H | 7684 | hCoV-19/Scotland/QEUH-96D220/2020\|EPI_ISL_530766\|2 | ···················· | ···················· | ···················· | ···················· | ···················· | ···················· |
| H | 7685 | hCoV-19/Scotland/QEUH-96D3C3/2020\|EPI_ISL_530767\|2 | ···················· | ···················· | ···················· | ···················· | ···················· | ···················· |
| H | 7686 | hCoV-19/Scotland/QEUH-96DDC5/2020\|EPI_ISL_530768\|2 | ···················· | ···················· | ···················· | ···················· | ···················· | ···················· |
| H | 7687 | hCoV-19/Northern Ireland/QEUH-96CBCC/2020\|EPI_ISL_ | ···················· | ···················· | ···················· | ···················· | ···················· | ···················· |
| H | 7688 | hCoV-19/Northern Ireland/QEUH-96DC13/2020\|EPI_ISL_ | ···················· | ···················· | ···················· | ···················· | ···················· | ···················· |
| H | 7689 | hCoV-19/Northern Ireland/QEUH-96CF93/2020\|EPI_ISL_ | ···················· | ···················· | ···················· | ···················· | ···················· | ···················· |
| H | 7690 | hCoV-19/England/QEUH-96DADD/2020\|EPI_ISL_530774\|20 | ···················· | ···················· | ···················· | ···················· | ···················· | ···············A···· |
| H | 7691 | hCoV-19/Northern Ireland/QEUH-96CB71/2020\|EPI_ISL_ | ···················· | ···················· | ···················· | ···················· | ···················· | ···················· |
| H | 7692 | hCoV-19/England/QEUH-96DA82/2020\|EPI_ISL_530776\|20 | ···················· | ···················· | ···················· | ···················· | ···················· | ···················· |
| H | 7693 | hCoV-19/Northern Ireland/QEUH-96D9C1/2020\|EPI_ISL_ | ···················· | ···················· | ···················· | ···················· | ···················· | ···················· |
| H | 7694 | hCoV-19/Northern Ireland/QEUH-96CBF9/2020\|EPI_ISL_ | ···················· | ···················· | ···················· | ···················· | ···················· | ···················· |
| H | 7695 | hCoV-19/Northern Ireland/QEUH-96CFB1/2020\|EPI_ISL_ | ···················· | ···················· | ···················· | ···················· | ···················· | ···················· |
| H | 7696 | hCoV-19/England/QEUH-96DEA4/2020\|EPI_ISL_530780\|20 | ···················· | ···················· | ···················· | ···················· | ···················· | ···················· |
| H | 7697 | hCoV-19/Northern Ireland/QEUH-96DBDA/2020\|EPI_ISL_ | ···················· | ···················· | ···················· | ···················· | ···················· | ···················· |
| H | 7698 | hCoV-19/Northern Ireland/QEUH-96CA56/2020\|EPI_ISL_ | ···················· | ···················· | ···················· | ···················· | ···················· | ···················· |
| H | 7699 | hCoV-19/Northern Ireland/QEUH-96CA65/2020\|EPI_ISL_ | ···················· | ···················· | ···················· | ···················· | ···················· | ···················· |
| H | 7700 | hCoV-19/Northern Ireland/QEUH-96CC9C/2020\|EPI_ISL_ | ···················· | ···················· | ···················· | ···················· | ···················· | ···················· |
| H | 7701 | hCoV-19/Northern Ireland/QEUH-96CACF/2020\|EPI_ISL_ | ···················· | ···················· | ···················· | ···················· | ···················· | ···················· |
| H | 7702 | hCoV-19/Northern Ireland/QEUH-96C9A4/2020\|EPI_ISL_ | ···················· | ···················· | ···················· | ···················· | ···················· | ···················· |
| H | 7703 | hCoV-19/England/QEUH-96C8F2/2020\|EPI_ISL_530788\|20 | ···················· | ···················· | ···················· | ···················· | ···················· | ···················· |
| H | 7704 | hCoV-19/Northern Ireland/QEUH-96DA46/2020\|EPI_ISL_ | ···················· | ···················· | ···················· | ···················· | ···················· | ···················· |
| H | 7705 | hCoV-19/Northern Ireland/QEUH-96D897/2020\|EPI_ISL_ | ···················· | ···················· | ···················· | ···················· | ···················· | ···················· |
| H | 7706 | hCoV-19/Northern Ireland/QEUH-96D5AF/2020\|EPI_ISL_ | ···················· | ···················· | ···················· | ···················· | ···················· | ···················· |
| H | 7707 | hCoV-19/England/QEUH-96E0AD/2020\|EPI_ISL_530793\|20 | ···················· | ···················· | ···················· | ···················· | ···················· | ···················· |
| H | 7708 | hCoV-19/Northern Ireland/QEUH-96DC6E/2020\|EPI_ISL_ | ···················· | ···················· | ···················· | ···················· | ···················· | ···················· |
| H | 7709 | hCoV-19/England/QEUH-96DAFB/2020\|EPI_ISL_530796\|20 | ···················· | ···················· | ···················· | ···················· | ···················· | ···················· |
| H | 7710 | hCoV-19/Northern Ireland/QEUH-96D0DB/2020\|EPI_ISL_ | ···················· | ···················· | ···················· | ···················· | ···················· | ···················· |
| H | 7711 | hCoV-19/England/QEUH-96C83E/2020\|EPI_ISL_530798\|20 | ···················· | ···················· | ···················· | ···················· | ···················· | ···················· |
| H | 7712 | hCoV-19/Latvia/111/2020\|EPI_ISL_534200\|2020-08-07 | ···················· | ···················· | ···················· | ···················· | ···················· | ···················· |
| H | 7713 | hCoV-19/Latvia/112/2020\|EPI_ISL_534201\|2020-08-10 | ···················· | ···················· | ···················· | ···················· | ···················· | ···················· |
| H | 7714 | hCoV-19/Latvia/113/2020\|EPI_ISL_534202\|2020-08-07 | ···················· | ···················· | ···················· | ···················· | ···················· | ···················· |
| H | 7715 | hCoV-19/Latvia/114/2020\|EPI_ISL_534203\|2020-08-08 | ···················· | ···················· | ···················· | ···················· | ···················· | ···················· |
| H | 7716 | hCoV-19/Latvia/115/2020\|EPI_ISL_534204\|2020-08-05 | ···················· | ···················· | ···················· | ···················· | ···················· | ···················· |
| H | 7717 | hCoV-19/Latvia/116/2020\|EPI_ISL_534205\|2020-08-06 | ···················· | ···················· | ···················· | ···················· | ···················· | ···················· |
| H | 7718 | hCoV-19/South Africa/KRISP-K002630/2020\|EPI_ISL_53 | ···················· | ···················· | ···················· | ···················· | ···················· | ···················· |
| H | 7719 | hCoV-19/South Africa/KRISP-K002631/2020\|EPI_ISL_53 | ···················· | ···················· | ···················· | ···················· | ···················· | ···················· |
| H | 7720 | hCoV-19/South Africa/KRISP-K002632/2020\|EPI_ISL_53 | ···················· | ···················· | ···················· | ···················· | ···················· | ···················· |
| H | 7721 | hCoV-19/South Africa/KRISP-K002633/2020\|EPI_ISL_53 | ···················· | ···················· | ···················· | ···················· | ···················· | ···················· |
| H | 7722 | hCoV-19/South Africa/KRISP-K002634/2020\|EPI_ISL_53 | ···················· | ···················· | ···················· | ···················· | ···················· | ···················· |
| H | 7723 | hCoV-19/South Africa/KRISP-K002635/2020\|EPI_ISL_53 | ···················· | ···················· | ···················· | ···················· | ····T··············· | ···················· |
| H | 7724 | hCoV-19/South Africa/KRISP-K002636/2020\|EPI_ISL_53 | ···················· | ···················· | ···················· | ···················· | ···················· | ···················· |
| H | 7725 | hCoV-19/South Africa/KRISP-K002637/2020\|EPI_ISL_53 | ···················· | ···················· | ···················· | ···················· | ···················· | ···················· |
| H | 7726 | hCoV-19/South Africa/KRISP-K002638/2020\|EPI_ISL_53 | ···················· | ···················· | ···················· | ···················· | ···················· | ···················· |
| H | 7727 | hCoV-19/South Africa/KRISP-K002639/2020\|EPI_ISL_53 | ···················· | ···················· | ···················· | ···················· | ···················· | ···················· |
| H | 7728 | hCoV-19/South Africa/KRISP-K002640/2020\|EPI_ISL_53 | ···················· | ···················· | ···················· | ···················· | ···················· | ···················· |
| H | 7729 | hCoV-19/South Africa/KRISP-K002641/2020\|EPI_ISL_53 | ···················· | ···················· | ···················· | ···················· | ···················· | ···················· |
| H | 7730 | hCoV-19/South Africa/KRISP-K002642/2020\|EPI_ISL_53 | ···················· | ···················· | ···················· | ···················· | ···················· | ···················· |
| H | 7731 | hCoV-19/South Africa/KRISP-K002643/2020\|EPI_ISL_53 | ···················· | ···················· | ···················· | ···················· | ···················· | ···················· |
| H | 7732 | hCoV-19/South Africa/KRISP-K002644/2020\|EPI_ISL_53 | ···················· | ···················· | ···················· | ···················· | ···················· | ···················· |
| H | 7733 | hCoV-19/South Africa/KRISP-K002645/2020\|EPI_ISL_53 | ···················· | ···················· | ···················· | ···················· | ···················· | ···················· |
| H | 7734 | hCoV-19/South Africa/KRISP-K002646/2020\|EPI_ISL_53 | ···················· | ···················· | ···················· | ···················· | ···················· | ···················· |
| H | 7735 | hCoV-19/South Africa/KRISP-K002648/2020\|EPI_ISL_53 | ···················· | ···················· | ···················· | ···················· | ···················· | ···················· |
| H | 7736 | hCoV-19/South Africa/KRISP-K002649/2020\|EPI_ISL_53 | ···················· | ···················· | ···················· | ···················· | ···················· | ···················· |
| H | 7737 | hCoV-19/South Africa/KRISP-K002650/2020\|EPI_ISL_53 | ···················· | ···················· | ···················· | ···················· | ···················· | ···················· |
| H | 7738 | hCoV-19/South Africa/KRISP-K002653/2020\|EPI_ISL_53 | ···················· | ···················· | ···················· | ···················· | ···················· | ···················· |
| H | 7739 | hCoV-19/South Africa/KRISP-K002655/2020\|EPI_ISL_53 | ···················· | ···················· | ···················· | ···················· | ···················· | ···················· |
| H | 7740 | hCoV-19/South Africa/KRISP-K002658/2020\|EPI_ISL_53 | ···················· | ···················· | ···················· | ···················· | ···················· | ···················· |
| H | 7741 | hCoV-19/South Africa/KRISP-K002659/2020\|EPI_ISL_53 | ···················· | ···················· | ···················· | ···················· | ···················· | ···················· |
| H | 7742 | hCoV-19/South Africa/KRISP-K002661/2020\|EPI_ISL_53 | ···················· | ···················· | ···················· | ···················· | ···················· | ···················· |
| H | 7743 | hCoV-19/South Africa/KRISP-K002664/2020\|EPI_ISL_53 | ···················· | ···················· | ···················· | ···················· | ···················· | ···················· |
| H | 7744 | hCoV-19/South Africa/KRISP-K002665/2020\|EPI_ISL_53 | ···················· | ···················· | ···················· | ···················· | ···················· | ···················· |
| H | 7745 | hCoV-19/South Africa/KRISP-K002666/2020\|EPI_ISL_53 | ···················· | ···················· | ···················· | ···················· | ···················· | ···················· |
| H | 7746 | hCoV-19/South Africa/KRISP-K002667/2020\|EPI_ISL_53 | ···················· | ···················· | ···················· | ···················· | ···················· | ···················· |
| H | 7747 | hCoV-19/South Africa/KRISP-K002668/2020\|EPI_ISL_53 | ···················· | ···················· | ···················· | ···················· | ···················· | ···················· |
| H | 7748 | hCoV-19/South Africa/KRISP-K002670/2020\|EPI_ISL_53 | ···················· | ···················· | ···················· | ···················· | ···················· | ···················· |
| H | 7749 | hCoV-19/England/QEUH-9644F6/2020\|EPI_ISL_536819\|20 | ···················· | ···················· | ···················· | ···················· | ···················· | ···················· |
| H | 7750 | hCoV-19/England/QEUH-96A1EA/2020\|EPI_ISL_536820\|20 | ···················· | ···················· | ···················· | ···················· | ···················· | ···················· |
| H | 7751 | hCoV-19/England/QEUH-9641C2/2020\|EPI_ISL_536821\|20 | ···················· | ···················· | ···················· | ···················· | ···················· | ···················· |
| H | 7752 | hCoV-19/Northern Ireland/QEUH-9650B5/2020\|EPI_ISL_ | ···················· | ···················· | ···················· | ···················· | ···················· | ···················· |
| H | 7753 | hCoV-19/England/QEUH-963AC8/2020\|EPI_ISL_536825\|20 | ···················· | ···················· | ···················· | ···················· | ···················· | ···················· |
| H | 7754 | hCoV-19/England/QEUH-96A37B/2020\|EPI_ISL_536826\|20 | ···················· | ···················· | ···················· | ···················· | ···················· | ···················· |
| H | 7755 | hCoV-19/Northern Ireland/QEUH-964414/2020\|EPI_ISL_ | ···················· | ···················· | ···················· | ···················· | ···················· | ···················· |
| H | 7756 | hCoV-19/England/QEUH-964432/2020\|EPI_ISL_536828\|20 | ···················· | ···················· | ···················· | ···················· | ···················· | ···················· |
| H | 7757 | hCoV-19/England/QEUH-963925/2020\|EPI_ISL_536829\|20 | ···················· | ···················· | ···················· | ···················· | ···················· | ···················· |
| H | 7758 | hCoV-19/Wales/QEUH-96A3F3/2020\|EPI_ISL_536830\|2020 | ···················· | ···················· | ···················· | ···················· | ···················· | ···················· |
| H | 7759 | hCoV-19/England/QEUH-964854/2020\|EPI_ISL_536831\|20 | ···················· | ···················· | ···················· | ···················· | ···················· | ···················· |
| H | 7760 | hCoV-19/England/QEUH-96457B/2020\|EPI_ISL_536833\|20 | ···················· | ···················· | ···················· | ···················· | ···················· | ···················· |
| H | 7761 | hCoV-19/Northern Ireland/QEUH-96412C/2020\|EPI_ISL_ | ···················· | ···················· | ···················· | ···················· | ···················· | ···················· |
| H | 7762 | hCoV-19/England/QEUH-9646C3/2020\|EPI_ISL_536837\|20 | ···················· | ···················· | ···················· | ···················· | ···················· | ···················· |
| H | 7763 | hCoV-19/England/QEUH-963F6F/2020\|EPI_ISL_536838\|20 | ···················· | ···················· | ···················· | ···················· | ···················· | ···················· |

| | | | | | | | | |
|---|---|---|---|---|---|---|---|---|
| H | 7764 hCoV-19/England/QEUH-964520/2020\|EPI_ISL_536839\|20 | ···················· | ···················· | ···················· | ···················· | ···················· | ···················· | |
| H | 7765 hCoV-19/England/QEUH-96398F/2020\|EPI_ISL_536840\|20 | ···················· | ···················· | ···················· | ···················· | ···················· | ···················· | |
| H | 7766 hCoV-19/England/QEUH-964371/2020\|EPI_ISL_536841\|20 | ···················· | ···················· | ···················· | ···················· | ···················· | ···················· | |
| H | 7767 hCoV-19/Northern Ireland/QEUH-96404D/2020\|EPI_ISL_ | ···················· | ···················· | ···················· | ···················· | ···················· | ···················· | |
| H | 7768 hCoV-19/Northern Ireland/QEUH-963EF9/2020\|EPI_ISL_ | ···················· | ···················· | ···················· | ···················· | ···················· | ···················· | |
| H | 7769 hCoV-19/England/QEUH-964283/2020\|EPI_ISL_536845\|20 | ···················· | ···················· | ···················· | ···················· | ···················· | ···················· | |
| H | 7770 hCoV-19/England/QEUH-96A3C6/2020\|EPI_ISL_536847\|20 | ···················· | ···················· | ···················· | ···················· | ···················· | ···················· | |
| H | 7771 hCoV-19/England/QEUH-96456C/2020\|EPI_ISL_536848\|20 | ···················· | ···················· | ···················· | ···················· | ···················· | ···················· | |
| H | 7772 hCoV-19/Northern Ireland/QEUH-96405C/2020\|EPI_ISL_ | ···················· | ···················· | ···················· | ···················· | ···················· | ···················· | |
| H | 7773 hCoV-19/England/QEUH-9647B1/2020\|EPI_ISL_536851\|20 | ···················· | ···················· | ···················· | ···················· | ···················· | ···················· | |
| H | 7774 hCoV-19/England/QEUH-9643CC/2020\|EPI_ISL_536852\|20 | ···················· | ···················· | ···················· | ···················· | ···················· | ···················· | |
| H | 7775 hCoV-19/England/QEUH-9648AF/2020\|EPI_ISL_536853\|20 | ···················· | ···················· | ···················· | ···················· | ···················· | ···················· | |
| H | 7776 hCoV-19/England/QEUH-9647DF/2020\|EPI_ISL_536854\|20 | ···················· | ···················· | ···················· | ···················· | ···················· | ···················· | |
| H | 7777 hCoV-19/England/QEUH-9646A5/2020\|EPI_ISL_536855\|20 | ···················· | ···················· | ···················· | ···················· | ···················· | ···················· | |
| H | 7778 hCoV-19/England/QEUH-9641E0/2020\|EPI_ISL_536857\|20 | ···················· | ·A·················· | ···················· | ···················· | ···················· | ··A················· | |
| H | 7779 hCoV-19/Northern Ireland/QEUH-964168/2020\|EPI_ISL_ | ···················· | ···················· | ···················· | ···················· | ···················· | ···················· | |
| H | 7780 hCoV-19/England/QEUH-9643BD/2020\|EPI_ISL_536861\|20 | ···················· | ···················· | ···················· | ···················· | ···················· | ···················· | |
| H | 7781 hCoV-19/England/QEUH-96465A/2020\|EPI_ISL_536862\|20 | ···················· | ···················· | ···················· | ···················· | ···················· | ···················· | |
| H | 7782 hCoV-19/England/QEUH-96A214/2020\|EPI_ISL_536863\|20 | ···················· | ···················· | ···················· | ···················· | ···················· | ···················· | |
| H | 7783 hCoV-19/England/QEUH-9647C0/2020\|EPI_ISL_536864\|20 | ···················· | ···················· | ···················· | ···················· | ···················· | ···················· | |
| H | 7784 hCoV-19/Northern Ireland/QEUH-964326/2020\|EPI_ISL_ | ···················· | ···················· | ···················· | ···················· | ···················· | ···················· | |
| H | 7785 hCoV-19/England/QEUH-96A320/2020\|EPI_ISL_536866\|20 | ···················· | ···················· | ···················· | ···················· | ···················· | ···················· | |
| H | 7786 hCoV-19/England/LEED-2A8B52/2020\|EPI_ISL_539074\|20 | ···················· | ···················· | ···················· | ···················· | ···················· | ···················· | |
| H | 7787 hCoV-19/England/LEED-2A8F83/2020\|EPI_ISL_539075\|20 | ···················· | ···················· | ···················· | ···················· | ···················· | ···················· | |
| H | 7788 hCoV-19/England/LEED-2A88B5/2020\|EPI_ISL_539076\|20 | ···················· | ···················· | ···················· | ···················· | ···················· | ···················· | |
| H | 7789 hCoV-19/England/LEED-2A8800/2020\|EPI_ISL_539077\|20 | ···················· | ···················A | ···················· | ···················· | ···················· | ···················· | 1 |
| H | 7790 hCoV-19/England/LEED-2A892B/2020\|EPI_ISL_539080\|20 | ···················· | ·············A······ | ···················· | ···················· | ···················· | ···················· | |
| H | 7791 hCoV-19/England/LEED-2A882E/2020\|EPI_ISL_539081\|20 | ···················· | ···················· | ···················· | ···················· | ···················· | ···················· | |
| H | 7792 hCoV-19/England/LEED-2A8BF8/2020\|EPI_ISL_539082\|20 | ···················· | ···················· | ···················· | ···················· | ···················· | ···················· | |
| H | 7793 hCoV-19/England/LEED-2A867F/2020\|EPI_ISL_539083\|20 | ···················· | ···················· | ···················· | ···················· | ···················· | ···················· | |
| H | 7794 hCoV-19/England/LEED-2A90AD/2020\|EPI_ISL_539084\|20 | ···················· | ···················A | ···················· | ···················· | ···················· | ···················· | 1 |
| H | 7795 hCoV-19/England/LEED-2A885B/2020\|EPI_ISL_539085\|20 | ···················· | ···················· | ···················· | ···················· | ···················· | ···················· | |
| H | 7796 hCoV-19/England/LEED-2A8606/2020\|EPI_ISL_539086\|20 | ···················· | ···················· | ···················· | ···················· | ···················· | ···················· | |
| H | 7797 hCoV-19/England/LEED-2A86CA/2020\|EPI_ISL_539087\|20 | ···················· | ·············A······ | ···················· | ···················· | ···················· | ···················· | |
| H | 7798 hCoV-19/England/LEED-2A8642/2020\|EPI_ISL_539089\|20 | ···················· | ···················· | ···················· | ···················· | ···················· | ···················· | |
| H | 7799 hCoV-19/England/LEED-2A8C5F/2020\|EPI_ISL_539090\|20 | ···················· | ·A·················· | ···················· | ···················· | ···················· | ···················· | |
| H | 7800 hCoV-19/England/LEED-2A90CB/2020\|EPI_ISL_539091\|20 | ···················· | ···················· | ···················· | ···················· | ···················· | ···················· | |
| H | 7801 hCoV-19/England/LEED-2A824E/2020\|EPI_ISL_539092\|20 | ···················· | ···················· | ···················· | ···················· | ···················· | ···················· | |
| H | 7802 hCoV-19/England/LEED-2A8FB0/2020\|EPI_ISL_539093\|20 | ···················· | ···················A | ···················· | ···················· | ···················· | ···················· | 1 |
| H | 7803 hCoV-19/England/LEED-2A8B61/2020\|EPI_ISL_539096\|20 | ···················· | ···················· | ···················· | ···················· | ···················· | ···················· | |
| H | 7804 hCoV-19/England/LEED-2A891C/2020\|EPI_ISL_539097\|20 | ···················· | ···················· | ···················· | ···················· | ···················· | ···················· | |
| H | 7805 hCoV-19/England/LEED-2A8D10/2020\|EPI_ISL_539099\|20 | ···················· | ···················· | ···················· | ···················· | ···················· | ···················· | |
| H | 7806 hCoV-19/England/LEED-2A85CD/2020\|EPI_ISL_539100\|20 | ···················· | ···················· | ···················· | ···················· | ···················· | ···················· | |
| H | 7807 hCoV-19/England/LEED-2A9016/2020\|EPI_ISL_539102\|20 | ···················· | ···················· | ···················· | ···················· | ···················· | ···················· | |
| H | 7808 hCoV-19/England/LEED-2A8ED1/2020\|EPI_ISL_539103\|20 | ···················· | ···················· | ···················· | ···················· | ···················· | ···················· | |
| H | 7809 hCoV-19/England/LEED-2A9043/2020\|EPI_ISL_539104\|20 | ···················· | ···················· | ···················· | ···················· | ···················· | ···················· | |
| H | 7810 hCoV-19/England/LEED-2A87B8/2020\|EPI_ISL_539106\|20 | ···················· | ···················· | ···················· | ···················· | ···················· | ···················· | |
| H | 7811 hCoV-19/England/LEED-2A8F92/2020\|EPI_ISL_539107\|20 | ···················· | ···················· | ···················· | ···················· | ···················· | ···················· | |
| H | 7812 hCoV-19/England/LEED-2A8FDE/2020\|EPI_ISL_539108\|20 | ···················· | ···················· | ···················· | ···················· | ···················· | ···················· | |
| H | 7813 hCoV-19/England/LEED-2A8A64/2020\|EPI_ISL_539109\|20 | ···················· | ···················· | ···················· | ···················· | ···················· | ···················· | |
| H | 7814 hCoV-19/England/LEED-2A88F1/2020\|EPI_ISL_539110\|20 | ···················· | ···················· | ···················· | ···················· | ···················· | ···················· | |
| H | 7815 hCoV-19/England/LEED-2A8B25/2020\|EPI_ISL_539111\|20 | ···················· | ···················· | ···················· | ···················· | ···················· | ···················· | |
| H | 7816 hCoV-19/England/LEED-2A89FE/2020\|EPI_ISL_539113\|20 | ···················· | ···················· | ···················· | ···················· | ···················· | ···················· | |
| H | 7817 hCoV-19/England/LEED-2A8CC8/2020\|EPI_ISL_539114\|20 | ···················· | ···················· | ···················· | ···················· | ···················· | ···················· | |
| H | 7818 hCoV-19/England/LEED-2A8C22/2020\|EPI_ISL_539115\|20 | ···················· | ···················· | ···················· | ···················· | ···················· | ···················· | |
| H | 7819 hCoV-19/England/LEED-2A8B16/2020\|EPI_ISL_539118\|20 | ···················· | ···················· | ···················· | ···················· | ···················· | ···················· | |
| H | 7820 hCoV-19/England/LEED-2A88C4/2020\|EPI_ISL_539119\|20 | ···················· | ···················· | ···················· | ···················· | ···················· | ···················· | |
| H | 7821 hCoV-19/England/LEED-2A876D/2020\|EPI_ISL_539121\|20 | ···················· | ···················· | ···················· | ···················· | ···················· | ···················· | |
| H | 7822 hCoV-19/England/LEED-2A8123/2020\|EPI_ISL_539122\|20 | ···················· | ···················· | ···················· | ···················· | ···················· | ···················· | |
| H | 7823 hCoV-19/England/LEED-2A8D5C/2020\|EPI_ISL_539123\|20 | ···················· | ·A·················· | ···················· | ···················· | ···················· | ···················· | |
| H | 7824 hCoV-19/England/LEED-2A8FCF/2020\|EPI_ISL_539124\|20 | ···················· | ···················A | ···················· | ···················· | ···················· | ···················· | 1 |
| H | 7825 hCoV-19/England/LEED-2A8FFC/2020\|EPI_ISL_539126\|20 | ···················· | ···················A | ···················· | ···················· | ···················· | ···················· | 1 |
| H | 7826 hCoV-19/England/LEED-2A8C13/2020\|EPI_ISL_539127\|20 | ···················· | ···················· | ···················· | ···················· | ···················· | ···················· | |
| H | 7827 hCoV-19/England/LEED-2A8C04/2020\|EPI_ISL_539128\|20 | ···················· | ···················· | ···················· | ···················· | ···················· | ···················· | |
| H | 7828 hCoV-19/England/LEED-2A879A/2020\|EPI_ISL_539129\|20 | ···················· | ···················· | ···················· | ···················· | ···················· | ···················· | |
| H | 7829 hCoV-19/England/QEUH-98FCFD/2020\|EPI_ISL_540060\|20 | ···················· | ···················· | ···················· | ···················· | ···················· | ···················· | |
| H | 7830 hCoV-19/England/QEUH-998A15/2020\|EPI_ISL_540061\|20 | ···················· | ···················· | ···················· | ···················· | ···················· | ···················· | |
| H | 7831 hCoV-19/Scotland/QEUH-98F8EA/2020\|EPI_ISL_540062\|2 | ···················· | ···················· | ···················· | ···················· | ···················· | ···················· | |
| H | 7832 hCoV-19/England/QEUH-990145/2020\|EPI_ISL_540063\|20 | ···················· | ···················· | ···················· | ···················· | ···················· | ···················· | |
| H | 7833 hCoV-19/England/QEUH-9901BE/2020\|EPI_ISL_540064\|20 | ···················· | ···················· | ···················· | ···················· | ···················· | ···················· | |
| H | 7834 hCoV-19/England/QEUH-9987A5/2020\|EPI_ISL_540066\|20 | ···················· | ···················· | ···················· | ···················· | ···················· | ···················· | |
| H | 7835 hCoV-19/England/QEUH-99864E/2020\|EPI_ISL_540069\|20 | ···················· | ···················· | ···················· | ···················· | ···················· | ···················· | |
| H | 7836 hCoV-19/England/QEUH-98FBB4/2020\|EPI_ISL_540070\|20 | ···················· | ···················· | ···················· | ···················· | ···················· | ···················· | |
| H | 7837 hCoV-19/England/QEUH-998CD3/2020\|EPI_ISL_540071\|20 | ···················· | ·A·················· | ···················· | ···················· | ···················· | ···················· | |
| H | 7838 hCoV-19/Scotland/QEUH-998A51/2020\|EPI_ISL_540072\|2 | ···················· | ···················· | ···················· | ···················· | ···················· | ···················· | |
| H | 7839 hCoV-19/England/QEUH-99044C/2020\|EPI_ISL_540074\|20 | ···················· | ···················· | ···················· | ···················· | ···················· | ···················· | |
| H | 7840 hCoV-19/England/QEUH-98FC84/2020\|EPI_ISL_540075\|20 | ···················· | ···················· | ···················· | ···················· | ···················· | ···················· | |
| H | 7841 hCoV-19/England/QEUH-9985C9/2020\|EPI_ISL_540076\|20 | ···················· | ···················· | ···················· | ···················· | ···················· | ···················· | |
| H | 7842 hCoV-19/England/QEUH-998875/2020\|EPI_ISL_540077\|20 | ···················· | ···················· | ···················· | ···················· | ···················· | ···················· | |
| H | 7843 hCoV-19/England/QEUH-99856F/2020\|EPI_ISL_540078\|20 | ···················· | ···················· | ···················· | ···················· | ···················· | ···················· | |

| | | | | | | | |
|---|---|---|---|---|---|---|---|
| H | 7844 hCoV-19/England/QEUH-998435/2020\|EPI_ISL_540079\|20 | .................... | .................... | .................... | .................... | .................... | .................... |
| H | 7845 hCoV-19/England/QEUH-98FF8B/2020\|EPI_ISL_540080\|20 | .................... | .................... | .................... | .................... | .................... | .................... |
| H | 7846 hCoV-19/England/QEUH-98F7B0/2020\|EPI_ISL_540081\|20 | .................... | .................... | .................... | .................... | ...T................ | .................... |
| H | 7847 hCoV-19/England/QEUH-998E46/2020\|EPI_ISL_540082\|20 | .................... | .A.................. | .................... | .................... | .................... | .................... |
| H | 7848 hCoV-19/England/QEUH-998990/2020\|EPI_ISL_540083\|20 | .................... | .A.................. | .................... | .................... | .................... | ..A................. |
| H | 7849 hCoV-19/England/QEUH-9989DC/2020\|EPI_ISL_540084\|20 | .................... | .................... | .................... | .................... | .................... | .................... |
| H | 7850 hCoV-19/England/QEUH-998884/2020\|EPI_ISL_540085\|20 | .................... | .A.................. | .................... | .................... | .................... | .................... |
| H | 7851 hCoV-19/England/QEUH-998C88/2020\|EPI_ISL_540086\|20 | .................... | .A.................. | .................... | .................... | .................... | .................... |
| H | 7852 hCoV-19/Northern Ireland/QEUH-998602/2020\|EPI_ISL_ | .................... | .................... | .................... | .................... | .................... | .................... |
| H | 7853 hCoV-19/England/QEUH-98FB87/2020\|EPI_ISL_540089\|20 | .................... | .................... | .................... | .................... | .................... | .................... |
| H | 7854 hCoV-19/England/QEUH-998356/2020\|EPI_ISL_540090\|20 | .................... | .................... | .................... | .................... | .................... | .................... |
| H | 7855 hCoV-19/England/QEUH-990628/2020\|EPI_ISL_540091\|20 | .................... | .................... | .................... | .................... | .................... | .................... |
| H | 7856 hCoV-19/England/QEUH-9900A2/2020\|EPI_ISL_540092\|20 | .................... | .A.................. | .................... | .................... | .................... | .................... |
| H | 7857 hCoV-19/England/QEUH-998778/2020\|EPI_ISL_540093\|20 | .................... | .................... | .................... | .................... | .................... | .................... |
| H | 7858 hCoV-19/England/QEUH-98FF5E/2020\|EPI_ISL_540094\|20 | .................... | .A.................. | .................... | .................... | .................... | .................... |
| H | 7859 hCoV-19/England/QEUH-98F48C/2020\|EPI_ISL_540095\|20 | .................... | .A.................. | .................... | .................... | .................... | .................... |
| H | 7860 hCoV-19/England/QEUH-99863F/2020\|EPI_ISL_540096\|20 | .................... | .................... | .................... | .................... | .................... | .................... |
| H | 7861 hCoV-19/England/QEUH-98FD45/2020\|EPI_ISL_540097\|20 | .................... | .................... | .................... | .................... | .................... | .................... |
| H | 7862 hCoV-19/England/QEUH-99037C/2020\|EPI_ISL_540098\|20 | .................... | .................... | .................... | .................... | .................... | .................... |
| H | 7863 hCoV-19/England/QEUH-9989FA/2020\|EPI_ISL_540101\|20 | .................... | .................... | .................... | .................... | .................... | .................... |
| H | 7864 hCoV-19/England/QEUH-998972/2020\|EPI_ISL_540102\|20 | .................... | .................... | .................... | .................... | .................... | .................... |
| H | 7865 hCoV-19/England/QEUH-98FBE1/2020\|EPI_ISL_540103\|20 | .................... | .................... | .................... | .................... | .................... | .................... |
| H | 7866 hCoV-19/England/QEUH-998918/2020\|EPI_ISL_540104\|20 | .................... | .................... | .................... | .................... | .................... | .................... |
| H | 7867 hCoV-19/England/QEUH-990233/2020\|EPI_ISL_540105\|20 | .................... | .................... | .................... | .................... | .................... | .................... |
| H | 7868 hCoV-19/England/QEUH-98FE24/2020\|EPI_ISL_540106\|20 | .................... | .................... | .................... | .................... | .................... | .................... |
| H | 7869 hCoV-19/England/QEUH-98F4AA/2020\|EPI_ISL_540107\|20 | .................... | .A.................. | .................... | .................... | .................... | .................... |
| H | 7870 hCoV-19/Scotland/QEUH-998787/2020\|EPI_ISL_540108\|2 | .................... | .................... | .................... | .................... | .................... | .................... |
| H | 7871 hCoV-19/England/QEUH-98FF7C/2020\|EPI_ISL_540109\|20 | .................... | .................... | .................... | .................... | .................... | .................... |
| H | 7872 hCoV-19/Scotland/QEUH-98F923/2020\|EPI_ISL_540110\|2 | .................... | .................... | .................... | .................... | .................... | .................... |
| H | 7873 hCoV-19/England/QEUH-998699/2020\|EPI_ISL_540112\|20 | .................... | .................... | .................... | .................... | .................... | .................... |
| H | 7874 hCoV-19/England/QEUH-998963/2020\|EPI_ISL_540119\|20 | .................... | .................... | .................... | .................... | .................... | .................... |
| H | 7875 hCoV-19/Brazil/SE-6591/2020\|EPI_ISL_541390\|2020-04 | .................... | .................... | .................... | .................... | .................... | .................... |
| H | 7876 hCoV-19/Brazil/SE-6594/2020\|EPI_ISL_541391\|2020-04 | .................... | .................... | .................... | .................... | .................... | .................... |
| H | 7877 hCoV-19/Brazil/SE-6603/2020\|EPI_ISL_541393\|2020-04 | .................... | .................... | .................... | .................... | .................... | .................... |
| H | 7878 hCoV-19/Brazil/SE-6606/2020\|EPI_ISL_541394\|2020-04 | .................... | .................... | .................... | .................... | .................... | .................... |
| H | 7879 hCoV-19/Brazil/SE-6607/2020\|EPI_ISL_541395\|2020-04 | .................... | .................... | .................... | .................... | .................... | .................... |
| H | 7880 hCoV-19/Switzerland/AG-ETHZ-270139/2020\|EPI_ISL_54 | .................... | .................... | .................... | .................... | .................... | .................... |
| H | 7881 hCoV-19/Switzerland/AG-ETHZ-270165/2020\|EPI_ISL_54 | .................... | .................... | .................... | .................... | .................... | .................... |
| H | 7882 hCoV-19/Switzerland/AG-ETHZ-270166/2020\|EPI_ISL_54 | .................... | .................... | .................... | .................... | .................... | .................... |
| H | 7883 hCoV-19/Switzerland/AG-ETHZ-270172/2020\|EPI_ISL_54 | .................... | .................... | .................... | .................... | .................... | .................... |
| H | 7884 hCoV-19/Switzerland/AG-ETHZ-270180/2020\|EPI_ISL_54 | .................... | .................... | .................... | .................... | .................... | .................... |
| H | 7885 hCoV-19/Switzerland/BE-ETHZ-270038/2020\|EPI_ISL_54 | .................... | .................... | .................... | .................... | .................... | .................... |
| H | 7886 hCoV-19/Switzerland/BE-ETHZ-270048/2020\|EPI_ISL_54 | .................... | .................... | .................... | .................... | .................... | .................... |
| H | 7887 hCoV-19/Switzerland/BE-ETHZ-270053/2020\|EPI_ISL_54 | .................... | .................... | .................... | .................... | .................... | .................... |
| H | 7888 hCoV-19/Switzerland/BE-ETHZ-270054/2020\|EPI_ISL_54 | .................... | .................... | .................... | .................... | .................... | .................... |
| H | 7889 hCoV-19/Switzerland/BE-ETHZ-270058/2020\|EPI_ISL_54 | .................... | .................... | .................... | .................... | .................... | .................... |
| H | 7890 hCoV-19/Switzerland/BE-ETHZ-270059/2020\|EPI_ISL_54 | .................... | .................... | .................... | .................... | .................... | .................... |
| H | 7891 hCoV-19/Switzerland/BE-ETHZ-270060/2020\|EPI_ISL_54 | .................... | .................... | .................... | .................... | .................... | .................... |
| H | 7892 hCoV-19/Switzerland/BE-ETHZ-270068/2020\|EPI_ISL_54 | .................... | .................... | .................... | .................... | .................... | .................... |
| H | 7893 hCoV-19/Switzerland/BE-ETHZ-270085/2020\|EPI_ISL_54 | .................... | .................... | .................... | .................... | .................... | .................... |
| H | 7894 hCoV-19/Switzerland/BE-ETHZ-270087/2020\|EPI_ISL_54 | .................... | .................... | .................... | .................... | .................... | .................... |
| H | 7895 hCoV-19/Switzerland/BE-ETHZ-270105/2020\|EPI_ISL_54 | .................... | .................... | .................... | .................... | .................... | .................... |
| H | 7896 hCoV-19/Switzerland/BE-ETHZ-270109/2020\|EPI_ISL_54 | .................... | .................... | .................... | .................... | .................... | .................... |
| H | 7897 hCoV-19/Switzerland/BE-ETHZ-270110/2020\|EPI_ISL_54 | .................... | .................... | .................... | .................... | .................... | .................... |
| H | 7898 hCoV-19/Switzerland/BE-ETHZ-270112/2020\|EPI_ISL_54 | .................... | .................... | .................... | .................... | .................... | .................... |
| H | 7899 hCoV-19/Switzerland/BE-ETHZ-270116/2020\|EPI_ISL_54 | .................... | .................... | .................... | .................... | .................... | .................... |
| H | 7900 hCoV-19/Switzerland/BE-ETHZ-270121/2020\|EPI_ISL_54 | .................... | .................... | .................... | .................... | .................... | .................... |
| H | 7901 hCoV-19/Switzerland/BE-ETHZ-270126/2020\|EPI_ISL_54 | .................... | .................... | .................... | .................... | .................... | .................... |
| H | 7902 hCoV-19/Switzerland/BE-ETHZ-270130/2020\|EPI_ISL_54 | .................... | .................... | .................... | .................... | .................... | .................... |
| H | 7903 hCoV-19/Switzerland/BE-ETHZ-270133/2020\|EPI_ISL_54 | .................... | .................... | .................... | .................... | ...............Y.... | .................... |
| H | 7904 hCoV-19/Switzerland/BE-ETHZ-270136/2020\|EPI_ISL_54 | .................... | .................... | .................... | .................... | .................... | .................... |
| H | 7905 hCoV-19/Switzerland/BE-ETHZ-270157/2020\|EPI_ISL_54 | .................... | .................... | .................... | .................... | .................... | .................... |
| H | 7906 hCoV-19/Switzerland/BE-ETHZ-270163/2020\|EPI_ISL_54 | .................... | .................... | .................... | .................... | .................... | .................... |
| H | 7907 hCoV-19/Switzerland/BE-ETHZ-270178/2020\|EPI_ISL_54 | .................... | .................... | .................... | .................... | .................... | .................... |
| H | 7908 hCoV-19/Switzerland/BE-ETHZ-270179/2020\|EPI_ISL_54 | .................... | .................... | .................... | .................... | .................... | .................... |
| H | 7909 hCoV-19/Switzerland/BE-ETHZ-270181/2020\|EPI_ISL_54 | .................... | .................... | .................... | .................... | .................... | .................... |
| H | 7910 hCoV-19/Switzerland/BE-ETHZ-270189/2020\|EPI_ISL_54 | .................... | .................... | .................... | .................... | .................... | .................... |
| H | 7911 hCoV-19/Switzerland/BE-ETHZ-270194/2020\|EPI_ISL_54 | .................... | .................... | .................... | .................... | .................... | .................... |
| H | 7912 hCoV-19/Switzerland/BL-ETHZ-270108/2020\|EPI_ISL_54 | .................... | .................... | .................... | .................... | .................... | .................... |
| H | 7913 hCoV-19/USA/TX-HMH-1478/2020\|EPI_ISL_542742\|2020-0 | .................... | .................... | .................... | .................... | .................... | .................... |
| H | 7914 hCoV-19/USA/TX-HMH-1481/2020\|EPI_ISL_542743\|2020-0 | .................... | .................... | .................... | .................... | .................... | .................... |
| H | 7915 hCoV-19/USA/TX-HMH-1482/2020\|EPI_ISL_542744\|2020-0 | .................... | .................... | .................... | .................... | .................... | .................... |
| H | 7916 hCoV-19/USA/TX-HMH-1484/2020\|EPI_ISL_542746\|2020-0 | .................... | .................... | .................... | .................... | .................... | .................... |
| H | 7917 hCoV-19/USA/TX-HMH-1486/2020\|EPI_ISL_542748\|2020-0 | .................... | .................... | .................... | .................... | .................... | .................... |
| H | 7918 hCoV-19/USA/TX-HMH-1487/2020\|EPI_ISL_542749\|2020-0 | .................... | .................... | .................... | .................... | .................... | .................... |
| H | 7919 hCoV-19/USA/TX-HMH-1490/2020\|EPI_ISL_542751\|2020-0 | .................... | .................... | .................... | .................... | .................... | .................... |
| H | 7920 hCoV-19/USA/TX-HMH-1492/2020\|EPI_ISL_542752\|2020-0 | .................... | .................... | .................... | .................... | .................... | .................... |
| H | 7921 hCoV-19/USA/TX-HMH-1495/2020\|EPI_ISL_542753\|2020-0 | .................... | .................... | .................... | .................... | .................... | .................... |
| H | 7923 hCoV-19/USA/TX-HMH-1497/2020\|EPI_ISL_542755\|2020-0 | .................... | .................... | .................... | .................... | .................... | .................... |
| H | 7924 hCoV-19/USA/TX-HMH-1499/2020\|EPI_ISL_542756\|2020-0 | .................... | .................... | .................... | .................... | .................... | .................... |

| | | | | | | | | | | |
|---|---|---|---|---|---|---|---|---|---|---|
| H | 7925 | hCoV-19/USA/TX-HMH-1500/2020\|EPI_ISL_542757\|2020-0 | ···················· | ···················· | ····················· | ···················· | ···················· | ···················· | | |
| H | 7926 | hCoV-19/USA/TX-HMH-1503/2020\|EPI_ISL_542759\|2020-0 | ···················· | ···················· | ····················· | ···················· | ···················· | ···················· | | |
| H | 7927 | hCoV-19/USA/TX-HMH-1505/2020\|EPI_ISL_542760\|2020-0 | ···················· | ···················· | ····················· | ···················· | ···················· | ···················· | | |
| H | 7929 | hCoV-19/USA/TX-HMH-1509/2020\|EPI_ISL_542763\|2020-0 | ···················· | ···················· | ····················· | ···················· | ···················· | ···················· | | |
| H | 7930 | hCoV-19/USA/TX-HMH-1515/2020\|EPI_ISL_542765\|2020-0 | ···················· | ···················· | ····················· | ···················· | ···················· | ···················· | | |
| H | 7931 | hCoV-19/USA/TX-HMH-1516/2020\|EPI_ISL_542766\|2020-0 | ···················· | ···················· | ····················· | ···················· | NNN················· | ···················· | | 1 |
| H | 7932 | hCoV-19/USA/TX-HMH-1518/2020\|EPI_ISL_542768\|2020-0 | ···················· | ···················· | ····················· | ···················· | ···················· | ···················· | | |
| H | 7933 | hCoV-19/USA/TX-HMH-1519/2020\|EPI_ISL_542769\|2020-0 | ···················· | ···················· | ····················· | ···················· | ···················· | ···················· | | |
| H | 7934 | hCoV-19/USA/TX-HMH-1521/2020\|EPI_ISL_542771\|2020-0 | ···················· | ···················· | ····················· | ···················· | NNN················· | ···················· | | 1 |
| H | 7935 | hCoV-19/USA/TX-HMH-1522/2020\|EPI_ISL_542772\|2020-0 | ···················· | ···················· | ····················· | ···················· | ···················· | ···················· | | |
| H | 7936 | hCoV-19/USA/TX-HMH-1524/2020\|EPI_ISL_542773\|2020-0 | ···················· | ···················· | ····················· | ···················· | ···················· | ···················· | | |
| H | 7937 | hCoV-19/Northern Ireland/QEUH-96DD3E/2020\|EPI_ISL_ | ···················· | ···················· | ····················· | ···················· | ···················· | ···················· | | |
| H | 7938 | hCoV-19/Northern Ireland/QEUH-96C9C2/2020\|EPI_ISL_ | ···················· | ···················· | ····················· | ···················· | ···················· | ···················· | | |
| H | 7939 | hCoV-19/England/QEUH-96D071/2020\|EPI_ISL_530702\|20 | ···················· | ···················· | ····················· | ···················· | ···················· | ···················· | | |
| H | 7940 | hCoV-19/Northern Ireland/QEUH-96DB9E/2020\|EPI_ISL_ | ···················· | ···················· | ····················· | ···················· | ···················· | ···················· | | |
| H | 7941 | hCoV-19/England/QEUH-96D080/2020\|EPI_ISL_530704\|20 | ···················· | ·R·················· | ····················· | ···················· | ···················· | ···················· | | |
| H | 7942 | hCoV-19/England/QEUH-96DC8C/2020\|EPI_ISL_530705\|20 | ···················· | ···················· | ····················· | ···················· | ···················· | ···················· | | |
| H | 7943 | hCoV-19/Northern Ireland/QEUH-96DB25/2020\|EPI_ISL_ | ···················· | ···················· | ····················· | ···················· | ···················· | ···················· | | |
| H | 7944 | hCoV-19/England/QEUH-96D017/2020\|EPI_ISL_530707\|20 | ···················· | ···················· | ····················· | ···················· | ······T············· | ···················· | | |
| H | 7945 | hCoV-19/Scotland/QEUH-96DD7A/2020\|EPI_ISL_530708\|2 | ···················· | ···················· | ····················· | ···················· | ···················· | ···················· | | |
| H | 7946 | hCoV-19/Northern Ireland/QEUH-96DC7D/2020\|EPI_ISL_ | ···················· | ···················· | ····················· | ···················· | ···················· | ···················· | | |
| H | 7947 | hCoV-19/Northern Ireland/QEUH-96DCC8/2020\|EPI_ISL_ | ···················· | ···················· | ····················· | ···················· | ···················· | ···················· | | |
| H | 7948 | hCoV-19/Northern Ireland/QEUH-96CA74/2020\|EPI_ISL_ | ···················· | ···················· | ····················· | ···················· | ···················· | ···················· | | |
| H | 7949 | hCoV-19/Northern Ireland/QEUH-96DC40/2020\|EPI_ISL_ | ···················· | ···················· | ····················· | ···················· | ···················· | ···················· | | |
| H | 7950 | hCoV-19/England/QEUH-96D123/2020\|EPI_ISL_530713\|20 | ···················· | ···················· | ····················· | ···················· | ···················· | ···················· | | |
| H | 7951 | hCoV-19/Northern Ireland/QEUH-96CA92/2020\|EPI_ISL_ | ···················· | ···················· | ····················· | ···················· | ···················· | ···················· | | |
| H | 7952 | hCoV-19/Northern Ireland/QEUH-96DC5F/2020\|EPI_ISL_ | ···················· | ···················· | ····················· | ···················· | ···················· | ···················· | | |
| H | 7953 | hCoV-19/England/QEUH-96C85C/2020\|EPI_ISL_530717\|20 | ···················· | ···················· | ····················· | ···················· | ···················· | ···················· | | |
| H | 7954 | hCoV-19/Northern Ireland/QEUH-96DB61/2020\|EPI_ISL_ | ···················· | ···A················ | ····················· | ···················· | ···················· | ···················· | | |
| H | 7955 | hCoV-19/England/QEUH-96C8B6/2020\|EPI_ISL_530719\|20 | ···················· | ···················· | ····················· | ···················· | ···················· | ···················· | | |
| H | 7956 | hCoV-19/Northern Ireland/QEUH-96DABF/2020\|EPI_ISL_ | ···················· | ···················· | ····················· | ···················· | ···················· | ···················· | | |
| H | 7957 | hCoV-19/Northern Ireland/QEUH-96CE96/2020\|EPI_ISL_ | ···················· | ···················· | ····················· | ···················· | ···················· | ···················· | | |
| H | 7958 | hCoV-19/Scotland/QEUH-96D2A8/2020\|EPI_ISL_530722\|2 | ···················· | ···················· | ····················· | ···················· | ···················· | ···················· | | |
| H | 7959 | hCoV-19/England/QEUH-96D7C7/2020\|EPI_ISL_530723\|20 | ···················· | ·A·················· | ····················· | ···················· | ···················· | ···················· | | |
| H | 7960 | hCoV-19/England/QEUH-96DCE6/2020\|EPI_ISL_530724\|20 | ···················· | ···················· | ····················· | ···················· | ···················· | ···················· | | |
| H | 7961 | hCoV-19/Northern Ireland/QEUH-96CAFC/2020\|EPI_ISL_ | ···················· | ···················· | ····················· | ···················· | ···················· | ···················· | | |
| H | 7962 | hCoV-19/England/QEUH-96DD98/2020\|EPI_ISL_530726\|20 | ···················· | ···················· | ····················· | ···················· | ···················· | ···················· | | |
| H | 7963 | hCoV-19/England/QEUH-96D84C/2020\|EPI_ISL_530727\|20 | ···················· | ···················· | ····················· | ···················· | ···················· | ···················· | | |
| H | 7964 | hCoV-19/Northern Ireland/QEUH-96DAA0/2020\|EPI_ISL_ | ···················· | ···················· | ····················· | ···················· | ···················· | ···················· | | |
| H | 7965 | hCoV-19/England/QEUH-96D439/2020\|EPI_ISL_530730\|20 | ···················· | ·A·················· | ····················· | ···················· | ···················· | ···················· | | |
| H | 7966 | hCoV-19/Northern Ireland/QEUH-96CA0B/2020\|EPI_ISL_ | ···················· | ···················· | ····················· | ···················· | ···················· | ···················· | | |
| H | 7967 | hCoV-19/Northern Ireland/QEUH-96CE78/2020\|EPI_ISL_ | ···················· | ···················· | ····················· | ···················· | ···················· | ···················· | | |
| H | 7968 | hCoV-19/Northern Ireland/QEUH-96DA19/2020\|EPI_ISL_ | ···················· | ···················· | ····················· | ···················· | ···················· | ···················· | | |
| H | 7969 | hCoV-19/Northern Ireland/QEUH-96DB52/2020\|EPI_ISL_ | ···················· | ···················· | ····················· | ···················· | ···················· | ···················· | | |
| H | 7970 | hCoV-19/Northern Ireland/QEUH-96CE4B/2020\|EPI_ISL_ | ···················· | ···················· | ····················· | ···················· | ···················· | ···················· | | |
| H | 7971 | hCoV-19/England/QEUH-96D141/2020\|EPI_ISL_530737\|20 | ···················· | ···················· | ····················· | ···················· | ···················· | ···················· | | |
| H | 7972 | hCoV-19/England/QEUH-96D202/2020\|EPI_ISL_530738\|20 | ···················· | ···················· | ····················· | ···················· | ···················· | ···················· | | |
| H | 7973 | hCoV-19/Northern Ireland/QEUH-96C995/2020\|EPI_ISL_ | ···················· | ···················· | ····················· | ···················· | ···················· | ···················· | | |
| H | 7974 | hCoV-19/Northern Ireland/QEUH-96CB44/2020\|EPI_ISL_ | ···················· | ···················· | ····················· | ···················· | ···················· | ···················· | | |
| H | 7975 | hCoV-19/Scotland/QEUH-96DE0E/2020\|EPI_ISL_530743\|2 | ···················· | ···················· | ····················· | ···················· | ···················· | ···················· | | |
| H | 7976 | hCoV-19/Northern Ireland/QEUH-96CE87/2020\|EPI_ISL_ | ···················· | ···················· | ····················· | ···················· | ···················· | ···················· | | |
| H | 7977 | hCoV-19/Northern Ireland/QEUH-96CAB0/2020\|EPI_ISL_ | ···················· | ···················· | ····················· | ···················· | ···················· | ···················· | | |
| H | 7978 | hCoV-19/England/QEUH-96DE59/2020\|EPI_ISL_530748\|20 | ···················· | ···················· | ····················· | ···················· | ···················· | ···················· | | |
| H | 7979 | hCoV-19/Northern Ireland/QEUH-96CADE/2020\|EPI_ISL_ | ···················· | ···················· | ····················· | ···················· | ···················· | ···················· | | |
| H | 7980 | hCoV-19/Northern Ireland/QEUH-96CBDB/2020\|EPI_ISL_ | ···················· | ···················· | ····················· | ···················· | ···················· | ···················· | | |
| H | 7981 | hCoV-19/Northern Ireland/QEUH-96DBF8/2020\|EPI_ISL_ | ···················· | ···················· | ····················· | ···················· | ···················· | ···················· | | |
| H | 7982 | hCoV-19/Northern Ireland/QEUH-96C9B3/2020\|EPI_ISL_ | ···················· | ···················· | ····················· | ···················· | ···················· | ···················· | | |
| H | 7983 | hCoV-19/Northern Ireland/QEUH-96CA47/2020\|EPI_ISL_ | ···················· | ···················· | ····················· | ···················· | ···················· | ···················· | | |
| H | 7984 | hCoV-19/Scotland/QEUH-96D1F6/2020\|EPI_ISL_530754\|2 | ···················· | ···················· | ····················· | ···················· | ···················· | ···················· | | |
| H | 7985 | hCoV-19/England/QEUH-96D2F3/2020\|EPI_ISL_530755\|20 | ···················· | ···················· | ····················· | ···················· | ···················· | ···················· | | |
| H | 7986 | hCoV-19/England/QEUH-96D5FA/2020\|EPI_ISL_530756\|20 | ···················· | ···················· | ····················· | ···················· | ······T············· | ···················· | | |
| H | 7987 | hCoV-19/England/QEUH-96D0EA/2020\|EPI_ISL_530758\|20 | ···················· | ···················· | ····················· | ···················· | ···················· | ···················· | | |
| H | 7988 | hCoV-19/Northern Ireland/QEUH-96C9FF/2020\|EPI_ISL_ | ···················· | ···················· | ····················· | ···················· | ···················· | ···················· | | |
| H | 7989 | hCoV-19/Scotland/QEUH-96CE2D/2020\|EPI_ISL_530760\|2 | ···················· | ···················· | ····················· | ···················· | ···················· | ···················· | | |
| H | 7990 | hCoV-19/South Africa/KRISP-K002548/2020\|EPI_ISL_53 | ···················· | ···················· | ····················· | ···················· | ···················· | ···················· | | |
| H | 7991 | hCoV-19/USA/IL-UW-1377/2020\|EPI_ISL_536748\|2020-09 | ···················· | ···················· | ····················· | ···················· | ···················· | ···················· | | |
| H | 7992 | hCoV-19/USA/WI-UW-1378/2020\|EPI_ISL_536749\|2020-09 | ···················· | ···················· | ····················· | ···················· | ···················· | ···················· | | |
| H | 7993 | hCoV-19/England/LEED-2A8C31/2020\|EPI_ISL_539130\|20 | ···················· | ···················· | ····················· | ···················· | ···················· | ···················· | | |
| H | 7994 | hCoV-19/England/LEED-2A8633/2020\|EPI_ISL_539131\|20 | ···················· | ···················· | ····················· | ···················· | ···················· | ···················· | | |
| H | 7995 | hCoV-19/England/LEED-2A8A46/2020\|EPI_ISL_539132\|20 | ···················· | ···················· | ····················· | ···················· | ···················· | ···················· | | |
| H | 7996 | hCoV-19/England/LEED-2A8FA1/2020\|EPI_ISL_539133\|20 | ···················· | ·A·················· | ····················· | ···················· | ···················· | ···················· | | |
| H | 7997 | hCoV-19/England/LEED-2A8CF5/2020\|EPI_ISL_539134\|20 | ···················· | ···················· | ····················· | ···················· | ···················· | ···················· | | |
| H | 7998 | hCoV-19/England/LEED-2A8CAA/2020\|EPI_ISL_539135\|20 | ···················· | ···················· | ····················· | ···················· | ···················· | ···················· | | |
| H | 7999 | hCoV-19/England/LEED-2A8879/2020\|EPI_ISL_539137\|20 | ···················· | ···················· | ····················· | ···················· | ···················· | ···················· | | |
| H | 8000 | hCoV-19/England/LEED-2A823F/2020\|EPI_ISL_539138\|20 | ···················· | ···················· | ····················· | ···················· | ···················· | ···················· | | |
| H | 8001 | hCoV-19/England/LEED-2A8F0B/2020\|EPI_ISL_539142\|20 | ···················· | ···················· | ····················· | ···················· | ···················· | ···················· | | |
| H | 8002 | hCoV-19/England/LEED-2A8CD7/2020\|EPI_ISL_539144\|20 | ···················· | ···················· | ····················· | ···················· | ···················· | ···················· | | |
| H | 8003 | hCoV-19/England/LEED-2A8994/2020\|EPI_ISL_539145\|20 | ···················· | ···A················ | ····················· | ···················· | ···················· | ···················· | | |
| H | 8004 | hCoV-19/England/LEED-2A9113/2020\|EPI_ISL_539146\|20 | ···················· | ···················A | ····················· | ···················· | ···················· | ···················· | 1 | |
| H | 8005 | hCoV-19/England/LEED-2A8D89/2020\|EPI_ISL_539147\|20 | ···················· | ···················· | ····················· | ···················· | ···················· | ···················· | | |

H 8006 hCoV-19/England/LEED-2A8D6B/2020|EPI_ISL_539148|20 .................... .................... .................... .................... .................... ....................
H 8007 hCoV-19/England/LEED-2A86E8/2020|EPI_ISL_539153|20 .................... ....A............... .................... .................... .................... ....................
H 8008 hCoV-19/England/LEED-2A8518/2020|EPI_ISL_539156|20 .................... .................... .................... .................... .................... ....................
H 8009 hCoV-19/England/LEED-2A8509/2020|EPI_ISL_539158|20 .................... .................... .................... .................... .................... ....................
H 8010 hCoV-19/England/LEED-2A8545/2020|EPI_ISL_539159|20 .................... .................... .................... .................... .................... ....................
H 8011 hCoV-19/England/LEED-2A8475/2020|EPI_ISL_539160|20 .................... .................... .................... .................... .................... ....................
H 8012 hCoV-19/England/LEED-2A8536/2020|EPI_ISL_539162|20 .................... .................... .................... .................... .................... ....................
H 8013 hCoV-19/England/LEED-2A8E68/2020|EPI_ISL_539163|20 .................... .................... .................... .................... .................... ....................
H 8014 hCoV-19/England/LEED-2A9395/2020|EPI_ISL_539165|20 .................... .................... .................... .................... .................... ....................
H 8015 hCoV-19/England/LEED-2A8457/2020|EPI_ISL_539166|20 .................... .................... .................... .................... .................... ....................
H 8016 hCoV-19/England/LEED-2A8DE3/2020|EPI_ISL_539167|20 .................... .................... .................... .................... .................... ....................
H 8017 hCoV-19/England/LEED-2A8484/2020|EPI_ISL_539169|20 .................... .................... .................... .................... .................... ....................
H 8018 hCoV-19/England/LEED-2A8712/2020|EPI_ISL_539170|20 .................... .................... .................... .................... .................... ....................
H 8019 hCoV-19/England/LEED-2A8563/2020|EPI_ISL_539171|20 .................... .................... .................... .................... .................... ....................
H 8020 hCoV-19/England/LEED-2A8554/2020|EPI_ISL_539172|20 .................... .................... .................... .................... .................... ....................
H 8021 hCoV-19/England/LEED-2A8703/2020|EPI_ISL_539173|20 .................... .................... .................... .................... .................... ....................
H 8022 hCoV-19/England/LEED-2A8581/2020|EPI_ISL_539174|20 .................... ....A............... .................... .................... .................... ....................
H 8023 hCoV-19/England/LEED-2A932C/2020|EPI_ISL_539175|20 .................... .A.................. .................... .................... .................... ....................
H 8024 hCoV-19/England/LEED-2A8E4A/2020|EPI_ISL_539176|20 .................... .................... .................... .................... .................... ....................
H 8025 hCoV-19/England/LEED-2A8E0E/2020|EPI_ISL_539178|20 .................... .................... .................... .................... .................... ....................
H 8026 hCoV-19/England/LEED-2A91F5/2020|EPI_ISL_539179|20 .................... .................... .................... .................... .................... ....................
H 8027 hCoV-19/England/QEUH-99866C/2020|EPI_ISL_540113|20 .................... .................... .................... .................... .................... ....................
H 8028 hCoV-19/England/QEUH-990206/2020|EPI_ISL_540115|20 .................... .................... .................... .................... .................... ....................
H 8029 hCoV-19/England/QEUH-998A60/2020|EPI_ISL_540116|20 .................... .................... .................... .................... .................... ....................
H 8030 hCoV-19/Scotland/QEUH-9989EB/2020|EPI_ISL_540117|2 .................... .................... .................... .................... .................... ....................
H 8031 hCoV-19/England/QEUH-998B6D/2020|EPI_ISL_540118|20 .................... .................... .................... .................... .................... ....................
H 8032 hCoV-19/England/QEUH-98FE7F/2020|EPI_ISL_540120|20 .................... .................... .................... .................... .................... ....................
H 8033 hCoV-19/Scotland/QEUH-990136/2020|EPI_ISL_540121|2 .................... .................... .................... .................... .................... ....................
H 8034 hCoV-19/England/QEUH-98F905/2020|EPI_ISL_540122|20 .................... .................... .................... .................... .................... ....................
H 8035 hCoV-19/England/QEUH-98F932/2020|EPI_ISL_540123|20 .................... .................... .................... .................... .................... ....................
H 8036 hCoV-19/England/QEUH-98F6E0/2020|EPI_ISL_540124|20 .................... .................... .................... .................... .................... ....................
H 8037 hCoV-19/England/QEUH-98F589/2020|EPI_ISL_540125|20 .................... .................... .................... .................... .................... ....................
H 8038 hCoV-19/England/QEUH-998417/2020|EPI_ISL_540126|20 .................... .................... .................... .................... .................... ....................
H 8039 hCoV-19/Northern Ireland/QEUH-990242/2020|EPI_ISL_ .................... .................... .................... .................... .................... ....................
H 8040 hCoV-19/England/QEUH-98F64A/2020|EPI_ISL_540128|20 .................... .................... .................... .................... .................... ....................
H 8041 hCoV-19/England/QEUH-99029D/2020|EPI_ISL_540129|20 .................... .................... .................... .................... .................... ....................
H 8042 hCoV-19/Scotland/QEUH-998AAC/2020|EPI_ISL_540130|2 .................... .................... .................... .................... .................... ....................
H 8043 hCoV-19/England/QEUH-998BC7/2020|EPI_ISL_540131|20 .................... .................... .................... .................... .................... ....................
H 8044 hCoV-19/England/QEUH-99039A/2020|EPI_ISL_540132|20 .................... .................... .................... .................... .................... ....................
H 8045 hCoV-19/Scotland/QEUH-98F7FC/2020|EPI_ISL_540134|2 .................... .................... .................... .................... .................... ....................
H 8046 hCoV-19/England/QEUH-998E19/2020|EPI_ISL_540135|20 .................... .A.................. .................... .................... .................... ....................
H 8047 hCoV-19/Wales/QEUH-998866/2020|EPI_ISL_540136|2020 .................... .................... .................... .................... .................... ....................
H 8048 hCoV-19/England/QEUH-998BE5/2020|EPI_ISL_540137|20 .................... .................... .................... .................... .................... ....................
H 8049 hCoV-19/Scotland/QEUH-998927/2020|EPI_ISL_540138|2 .................... .................... .................... .................... .................... ....................
H 8050 hCoV-19/England/QEUH-9A43C0/2020|EPI_ISL_540139|20 .................... .A.......A.......... .................... .................... .................... ....................
H 8051 hCoV-19/England/QEUH-9A4278/2020|EPI_ISL_540140|20 .................... .A.................. .................... .................... .................... ....................
H 8052 hCoV-19/England/QEUH-9A4533/2020|EPI_ISL_540141|20 .................... .................... .................... .................... .................... ....................
H 8053 hCoV-19/England/QEUH-9A42F0/2020|EPI_ISL_540142|20 .................... .................... .................... .................... .................... ....................
H 8054 hCoV-19/England/QEUH-9A4490/2020|EPI_ISL_540143|20 .................... .A.................. .................... .................... .................... ....................
H 8055 hCoV-19/England/QEUH-9A4603/2020|EPI_ISL_540144|20 .................... .................... .................... .................... .................... ....................
H 8056 hCoV-19/England/QEUH-9A42B4/2020|EPI_ISL_540145|20 .................... .................... .................... .................... .................... ....................
H 8057 hCoV-19/England/QEUH-9A40E7/2020|EPI_ISL_540146|20 .................... .................... .................... .................... .................... ....................
H 8058 hCoV-19/England/QEUH-9A413F/2020|EPI_ISL_540147|20 .................... .................... .................... .................... .................... ....................
H 8059 hCoV-19/England/QEUH-9A4005/2020|EPI_ISL_540148|20 .................... .................... .................... .................... .................... ....................
H 8060 hCoV-19/England/QEUH-9A4427/2020|EPI_ISL_540149|20 .................... .................... .................... .................... ....T............... ....................
H 8061 hCoV-19/England/QEUH-9A3F54/2020|EPI_ISL_540150|20 .................... .................... .................... .................... .................... ....................
H 8062 hCoV-19/England/QEUH-9A42C3/2020|EPI_ISL_540151|20 .................... .................... .................... .................... .................... ....................
H 8063 hCoV-19/England/QEUH-9A45CA/2020|EPI_ISL_540152|20 .................... .................... .................... .................... .................... ....................
H 8064 hCoV-19/England/QEUH-9A40BA/2020|EPI_ISL_540154|20 .................... .................... .................... .................... .................... ....................
H 8065 hCoV-19/England/QEUH-9A425A/2020|EPI_ISL_540155|20 .................... .................... .................... .................... .................... ....................
H 8066 hCoV-19/England/QEUH-9A43FD/2020|EPI_ISL_540157|20 .................... .................... .................... .................... .................... ....................
H 8067 hCoV-19/England/QEUH-9A406F/2020|EPI_ISL_540159|20 .................... .................... .................... .................... .................... ....................
H 8068 hCoV-19/England/QEUH-9A467C/2020|EPI_ISL_540160|20 .................... .................... .................... .................... .................... ....................
H 8069 hCoV-19/England/QEUH-9A41C6/2020|EPI_ISL_540162|20 .................... .A.................. .................... .................... .................... ....................
H 8070 hCoV-19/England/QEUH-9A42E1/2020|EPI_ISL_540163|20 .................... .................... .................... .................... .................... ....................
H 8071 hCoV-19/England/QEUH-9A40D8/2020|EPI_ISL_540164|20 .................... .................... .................... .................... .................... ....................
H 8072 hCoV-19/England/QEUH-9A43DF/2020|EPI_ISL_540166|20 .................... .A.................. .................... .................... .................... ....................
H 8073 hCoV-19/England/QEUH-9A4023/2020|EPI_ISL_540167|20 .................... .................... .................... .................... .................... ....................
H 8074 hCoV-19/Switzerland/AG-ETHZ-270037/2020|EPI_ISL_54 .................... .................... .................... .................... ......K............. ....................
H 8075 hCoV-19/Switzerland/AG-ETHZ-270046/2020|EPI_ISL_54 .................... .................... .................... .................... .................... ....................
H 8076 hCoV-19/Switzerland/AG-ETHZ-270067/2020|EPI_ISL_54 .................... .................... .................... .................... ......K............. ....................
H 8077 hCoV-19/Switzerland/AG-ETHZ-270088/2020|EPI_ISL_54 .................... .................... .................... .................... ......K............. ....................
H 8078 hCoV-19/Switzerland/AG-ETHZ-270091/2020|EPI_ISL_54 .................... ....G............... .................... .................... .................... ....................
H 8079 hCoV-19/Switzerland/AG-ETHZ-270106/2020|EPI_ISL_54 .................... .................... .................... .................... ............R....... .....Y..............
H 8080 hCoV-19/Switzerland/AG-ETHZ-270107/2020|EPI_ISL_54 .................... .................... .................... .................... .................... ....................
H 8081 hCoV-19/Switzerland/AG-ETHZ-270111/2020|EPI_ISL_54 .................... .................... .................... .................... ......K............. ....................
H 8082 hCoV-19/Switzerland/AG-ETHZ-270120/2020|EPI_ISL_54 ........T........... .................... .................... .................... .................... ....................
H 8083 hCoV-19/Switzerland/BE-ETHZ-270203/2020|EPI_ISL_54 .................... .................... .................... .................... .................... ....................
H 8084 hCoV-19/Switzerland/BE-ETHZ-270209/2020|EPI_ISL_54 .................... .................... .................... .................... .................... ....................
H 8085 hCoV-19/Switzerland/BE-ETHZ-270218/2020|EPI_ISL_54 .................... .................... .................... .................... .................... ....................

H 8086 hCoV-19/Switzerland/BL-ETHZ-270079/2020|EPI_ISL_54 ···················· ···················· ···················· ···················· ···················· ····················
H 8087 hCoV-19/Switzerland/BL-ETHZ-270101/2020|EPI_ISL_54 ···················· ····G··············· ···················· ···················· ···················· ····················
H 8088 hCoV-19/Switzerland/BL-ETHZ-270134/2020|EPI_ISL_54 ···················· ···················· ···················· ···················· ···················· ····················
H 8089 hCoV-19/Switzerland/BL-ETHZ-270140/2020|EPI_ISL_54 ···················· ···················· ···················· ···················· ···················· ····················
H 8090 hCoV-19/Switzerland/BL-ETHZ-270149/2020|EPI_ISL_54 ···················· ···················· ···················· ···················· ···················· ····················
H 8091 hCoV-19/Switzerland/BL-ETHZ-270150/2020|EPI_ISL_54 ···················· ···················· ···················· ···················· ···················· ····················
H 8092 hCoV-19/Switzerland/BL-ETHZ-270153/2020|EPI_ISL_54 ···················· ···················· ···················· ···················· ···················· ····················
H 8093 hCoV-19/Switzerland/BL-ETHZ-270159/2020|EPI_ISL_54 ···················· ···················· ···················· ···················· ···················· ····················
H 8094 hCoV-19/Switzerland/BL-ETHZ-270160/2020|EPI_ISL_54 ···················· ···················· ···················· ···················· ···················· ····················
H 8095 hCoV-19/Switzerland/BL-ETHZ-270171/2020|EPI_ISL_54 ···················· ···················· ···················· ···················· ···················· ····················
H 8096 hCoV-19/Switzerland/BL-ETHZ-270210/2020|EPI_ISL_54 ···················· ···················· ···················· ···················· ···················· ····················
H 8097 hCoV-19/Switzerland/BL-ETHZ-270213/2020|EPI_ISL_54 ···················· ···················· ···················· ···················· ···················· ····················
H 8098 hCoV-19/Switzerland/BL-ETHZ-270214/2020|EPI_ISL_54 ···················· ···················· ···················· ···················· ···················· ····················
H 8099 hCoV-19/Switzerland/BL-ETHZ-270215/2020|EPI_ISL_54 ···················· ···················· ···················· ···················· ···················· ····················
H 8100 hCoV-19/Switzerland/BL-ETHZ-270216/2020|EPI_ISL_54 ···················· ···················· ···················· ···················· ···················· ····················
H 8101 hCoV-19/Switzerland/BS-ETHZ-270197/2020|EPI_ISL_54 ···················· ···················· ···················· ···················· ···················· ····················
H 8102 hCoV-19/Switzerland/GE-ETHZ-270137/2020|EPI_ISL_54 ···················· ···················· ···················· ···················· ···················· ····················
H 8103 hCoV-19/Switzerland/GR-ETHZ-270132/2020|EPI_ISL_54 ···················· ···················· ···················· ···················· ···················· ····················
H 8104 hCoV-19/Switzerland/LU-ETHZ-270042/2020|EPI_ISL_54 ···················· ···················· ···················· ···················· ···················· ····················
H 8105 hCoV-19/Switzerland/LU-ETHZ-270082/2020|EPI_ISL_54 ···················· ···················· ···················· ···················· ···················· ····················
H 8106 hCoV-19/Switzerland/SG-ETHZ-270043/2020|EPI_ISL_54 ···················· ···················· ···················· ···················· ···················· ····················
H 8107 hCoV-19/Switzerland/SG-ETHZ-270204/2020|EPI_ISL_54 ···················· ···················· ···················· ···················· ···················· ····················
H 8108 hCoV-19/Switzerland/VD-ETHZ-270050/2020|EPI_ISL_54 ···················· ···················· ···················· ···················· ···················· ····················
H 8109 hCoV-19/Switzerland/VD-ETHZ-270052/2020|EPI_ISL_54 ···················· ···················· ···················· ···················· ·······S············ ····················
H 8110 hCoV-19/Switzerland/VD-ETHZ-270056/2020|EPI_ISL_54 ···················· ···················· ···················· ···················· ···················· ····················
H 8111 hCoV-19/Switzerland/VD-ETHZ-270064/2020|EPI_ISL_54 ···················· ···················· ···················· ···················· ···················· ····················
H 8112 hCoV-19/Switzerland/VD-ETHZ-270065/2020|EPI_ISL_54 ···················· ···················· ···················· ···················· ···················· ····················
H 8113 hCoV-19/Switzerland/VD-ETHZ-270066/2020|EPI_ISL_54 ···················· ···················· ···················· ···················· ···················· ····················
H 8114 hCoV-19/Switzerland/VD-ETHZ-270072/2020|EPI_ISL_54 ···················· ···················· ···················· ···················· ···················· ·········Y··········
H 8115 hCoV-19/Switzerland/VD-ETHZ-270075/2020|EPI_ISL_54 ···················· ···················· ···················· ···················· ···················· ········R···········
H 8116 hCoV-19/Switzerland/VD-ETHZ-270076/2020|EPI_ISL_54 ···················· ···················· ···················· ···················· ···················· ····················
H 8117 hCoV-19/Switzerland/VD-ETHZ-270077/2020|EPI_ISL_54 ···················· ···················· ···················· ···················· ·······S············ ····················
H 8118 hCoV-19/Switzerland/VD-ETHZ-270080/2020|EPI_ISL_54 ···················· ···················· ···················· ···················· ···················· ····················
H 8119 hCoV-19/Switzerland/VD-ETHZ-270095/2020|EPI_ISL_54 ···················· ···················· ···················· ···················· ···················· ····················
H 8120 hCoV-19/Switzerland/VD-ETHZ-270097/2020|EPI_ISL_54 ···················· ···················· ···················· ···················· ···················· ····················
H 8121 hCoV-19/Switzerland/VD-ETHZ-270098/2020|EPI_ISL_54 ···················· ···················· ···················· ···················· ···················· ····················
H 8122 hCoV-19/Switzerland/VD-ETHZ-270102/2020|EPI_ISL_54 ···················· ···················· ···················· ···················· ···················· ····················
H 8123 hCoV-19/Switzerland/VD-ETHZ-270103/2020|EPI_ISL_54 ············T······· ···················· ···················· ···················· ···················· ····················
H 8124 hCoV-19/Switzerland/VD-ETHZ-270113/2020|EPI_ISL_54 ···················· ···················· ···················· ···················· ···················· ····················
H 8125 hCoV-19/Switzerland/VD-ETHZ-270114/2020|EPI_ISL_54 ···················· ···················· ···················· ···················· ···················· ····················
H 8126 hCoV-19/Switzerland/VD-ETHZ-270115/2020|EPI_ISL_54 ···················· ···················· ···················· ···················· ················M··· ····················
H 8127 hCoV-19/Switzerland/VD-ETHZ-270122/2020|EPI_ISL_54 ···················· ···················· ···················· ···················· ···················· ····················
H 8128 hCoV-19/Switzerland/VD-ETHZ-270123/2020|EPI_ISL_54 ···················· ···················· ···················· ···················· ·······S············ ····················
H 8129 hCoV-19/Switzerland/VD-ETHZ-270124/2020|EPI_ISL_54 ···················· ···················· ···················· ···················· ···················· ····················
H 8130 hCoV-19/Switzerland/VD-ETHZ-270125/2020|EPI_ISL_54 ···················· ···················· ···················· ···················· ···················· ····················
H 8131 hCoV-19/Switzerland/VD-ETHZ-270129/2020|EPI_ISL_54 ···················· ···················· ···················· ···················· ··········Y········· ····················
H 8132 hCoV-19/Switzerland/VD-ETHZ-270169/2020|EPI_ISL_54 ···················· ···················· ···················· ···················· ···················· ····················
H 8133 hCoV-19/Switzerland/VD-ETHZ-270174/2020|EPI_ISL_54 ···················· ···················· ···················· ···················· ···················· ····················
H 8134 hCoV-19/USA/TX-HMH-1402/2020|EPI_ISL_542700|2020-0 ···················· ···················· ···················· ···················· ···················· ····················
H 8135 hCoV-19/USA/TX-HMH-1403/2020|EPI_ISL_542701|2020-0 ···················· ···················· ···················· ···················· ···················· ····················
H 8136 hCoV-19/USA/TX-HMH-1404/2020|EPI_ISL_542702|2020-0 ···················· ···················· ···················· ···················· ···················· ····················
H 8137 hCoV-19/USA/TX-HMH-1405/2020|EPI_ISL_542703|2020-0 ···················· ···················· ···················· ···················· ···················· ····················
H 8138 hCoV-19/USA/TX-HMH-1406/2020|EPI_ISL_542704|2020-0 ···················· ···················· ···················· ···················· ···················· ····················
H 8139 hCoV-19/USA/TX-HMH-1408/2020|EPI_ISL_542706|2020-0 ···················· ···················· ···················· ···················· ···················· ····················
H 8140 hCoV-19/USA/TX-HMH-1409/2020|EPI_ISL_542707|2020-0 ···················· ··········A········· ···················· ···················· ···················· ····················
H 8141 hCoV-19/USA/TX-HMH-1410/2020|EPI_ISL_542708|2020-0 ···················· ···················· ···················· ···················· ···················· ····················
H 8142 hCoV-19/USA/TX-HMH-1415/2020|EPI_ISL_542709|2020-0 ···················· ···················· ·············T······ ···················· ···················· ····················
H 8143 hCoV-19/USA/TX-HMH-1418/2020|EPI_ISL_542711|2020-0 ···················· ···················· ···················· ···················· ···················· ····················
H 8144 hCoV-19/USA/TX-HMH-1419/2020|EPI_ISL_542712|2020-0 ···················· ···················· ···················· ···················· ···················· ····················
H 8145 hCoV-19/USA/TX-HMH-1421/2020|EPI_ISL_542713|2020-0 ···················· ···················· ···················· ···················· ···················· ····················
H 8146 hCoV-19/USA/TX-HMH-1422/2020|EPI_ISL_542714|2020-0 ···················· ···················· ···················· ···················· ···················· ····················
H 8147 hCoV-19/USA/TX-HMH-1424/2020|EPI_ISL_542715|2020-0 ···················· ···················· ···················· ···················· ···················· ····················
H 8148 hCoV-19/USA/TX-HMH-1425/2020|EPI_ISL_542716|2020-0 ···················· ···················· ···················· ···················· ···················· ····················
H 8149 hCoV-19/USA/TX-HMH-1426/2020|EPI_ISL_542717|2020-0 ···················· ···················· ···················· ···················· ···················· ····················
H 8150 hCoV-19/USA/TX-HMH-1427/2020|EPI_ISL_542718|2020-0 ···················· ···················· ···················· ···················· ···················· ····················
H 8151 hCoV-19/USA/TX-HMH-1428/2020|EPI_ISL_542719|2020-0 ···················· ···················· ···················· ···················· ···················· ····················
H 8153 hCoV-19/USA/TX-HMH-1433/2020|EPI_ISL_542721|2020-0 ···················· ···················· ···················· ···················· ···················· ····················
H 8154 hCoV-19/USA/TX-HMH-1435/2020|EPI_ISL_542722|2020-0 ···················· ···················· ···················· ···················· ···················· ····················
H 8155 hCoV-19/USA/TX-HMH-1436/2020|EPI_ISL_542723|2020-0 ···················· ···················· ···················· ···················· ···················· ····················
H 8156 hCoV-19/USA/TX-HMH-1439/2020|EPI_ISL_542724|2020-0 ···················· ···················· ···················· ···················· ···················· ····················
H 8157 hCoV-19/USA/TX-HMH-1441/2020|EPI_ISL_542725|2020-0 ···················· ···················· ···················· ···················· ···················· ····················
H 8158 hCoV-19/USA/TX-HMH-1443/2020|EPI_ISL_542726|2020-0 ···················· ···················· ···················· ···················· ···················· ····················
H 8159 hCoV-19/USA/TX-HMH-1444/2020|EPI_ISL_542727|2020-0 ···················· ···················· ···················· ···················· ···················· ····················
H 8160 hCoV-19/USA/TX-HMH-1445/2020|EPI_ISL_542728|2020-0 ···················· ···················· ···················· ···················· ···················· ····················
H 8161 hCoV-19/USA/TX-HMH-1449/2020|EPI_ISL_542729|2020-0 ···················· ···················· ···················· ···················· ···················· ····················
H 8162 hCoV-19/USA/TX-HMH-1452/2020|EPI_ISL_542730|2020-0 ···················· ···················· ···················· ···················· ···················· ····················
H 8163 hCoV-19/USA/TX-HMH-1453/2020|EPI_ISL_542731|2020-0 ···················· ···················· ···················· ···················· ···················· ····················
H 8164 hCoV-19/USA/TX-HMH-1458/2020|EPI_ISL_542733|2020-0 ···················· ···················· ···················· ···················· ···················· ····················
H 8166 hCoV-19/USA/TX-HMH-1461/2020|EPI_ISL_542735|2020-0 ···················· ···················· ···················· ···················· ···················· ····················
H 8167 hCoV-19/USA/TX-HMH-1464/2020|EPI_ISL_542736|2020-0 ···················· ···················· ···················· ···················· ···················· ····················

| | | | | | | | | |
|---|---|---|---|---|---|---|---|---|
| H | 8168 | hCoV-19/USA/TX-HMH-1466/2020\|EPI_ISL_542738\|2020-0 | ···················· | ···················· | ····················· | ···················· | ···················· | ···················· |
| H | 8169 | hCoV-19/USA/TX-HMH-1467/2020\|EPI_ISL_542739\|2020-0 | ···················· | ···················· | ····················· | ···················· | ···················· | ···················· |
| H | 8170 | hCoV-19/USA/TX-HMH-1468/2020\|EPI_ISL_542740\|2020-0 | ···················· | ···················· | ····················· | ···················· | ···················· | ···················· |
| H | 8171 | hCoV-19/USA/TX-HMH-1528/2020\|EPI_ISL_542775\|2020-0 | ···················· | ···················· | ····················· | ···················· | ···················· | ···················· |
| H | 8172 | hCoV-19/USA/TX-HMH-1529/2020\|EPI_ISL_542776\|2020-0 | ···················· | ···················· | ····················· | ···················· | ···················· | ···················· |
| H | 8173 | hCoV-19/USA/TX-HMH-1530/2020\|EPI_ISL_542777\|2020-0 | ···················· | ···················· | ····················· | ···················· | ···················· | ···················· |
| H | 8174 | hCoV-19/USA/TX-HMH-1531/2020\|EPI_ISL_542778\|2020-0 | ···················· | ···················· | ····················· | ···················· | ···················· | ···················· |
| H | 8175 | hCoV-19/USA/TX-HMH-1537/2020\|EPI_ISL_542779\|2020-0 | ···················· | ···················· | ····················· | ···················· | ···················· | ···················· |
| H | 8176 | hCoV-19/USA/TX-HMH-1539/2020\|EPI_ISL_542780\|2020-0 | ···················· | ···················· | ····················· | ···················· | ···················· | ···················· |
| H | 8177 | hCoV-19/USA/TX-HMH-1542/2020\|EPI_ISL_542782\|2020-0 | ···················· | ···················· | ····················· | ···················· | ···················· | ···················· |
| H | 8178 | hCoV-19/USA/TX-HMH-1544/2020\|EPI_ISL_542784\|2020-0 | ···················· | ···················· | ····················· | ···················· | ···················· | ···················· |
| H | 8179 | hCoV-19/USA/TX-HMH-1545/2020\|EPI_ISL_542785\|2020-0 | ···················· | ···················· | ····················· | ···················· | ···················· | ···················· |
| H | 8180 | hCoV-19/USA/TX-HMH-1546/2020\|EPI_ISL_542786\|2020-0 | ···················· | ···················· | ····················· | ···················· | ···················· | ···················· |
| H | 8181 | hCoV-19/USA/TX-HMH-1547/2020\|EPI_ISL_542787\|2020-0 | ···················· | ···················· | ····················· | ···················· | ···················· | ···················· |
| H | 8182 | hCoV-19/USA/TX-HMH-1548/2020\|EPI_ISL_542788\|2020-0 | ···················· | ···················· | ····················· | ···················· | ··············N····· | ·······N············ |
| H | 8183 | hCoV-19/USA/TX-HMH-1719/2020\|EPI_ISL_542789\|2020-0 | ···················· | ···················· | ····················· | ···················· | ···················· | ···················· |
| H | 8184 | hCoV-19/USA/TX-HMH-1721/2020\|EPI_ISL_542790\|2020-0 | ···················· | ···················· | ····················· | ···················· | ···················· | ···················· |
| H | 8185 | hCoV-19/USA/TX-HMH-1722/2020\|EPI_ISL_542791\|2020-0 | ···················· | ···················· | ····················· | ···················· | ···················· | ···················· |
| H | 8186 | hCoV-19/USA/TX-HMH-1723/2020\|EPI_ISL_542792\|2020-0 | ········T··········· | ···················· | ····················· | ···················· | ···················· | ···················· |
| H | 8187 | hCoV-19/USA/TX-HMH-1725/2020\|EPI_ISL_542794\|2020-0 | ···················· | ···················· | ····················· | ···················· | ···················· | ···················· |
| H | 8188 | hCoV-19/USA/TX-HMH-1726/2020\|EPI_ISL_542795\|2020-0 | ···················· | ···················· | ····················· | ···················· | ···················· | ···················· |
| H | 8189 | hCoV-19/USA/TX-HMH-1727/2020\|EPI_ISL_542796\|2020-0 | ···················· | ···················· | ····················· | ···················· | ···················· | ···················· |
| H | 8190 | hCoV-19/USA/TX-HMH-1728/2020\|EPI_ISL_542797\|2020-0 | ···················· | ··············C····· | ····················· | ···················· | ···················· | ···················· |
| H | 8191 | hCoV-19/USA/TX-HMH-1729/2020\|EPI_ISL_542798\|2020-0 | ···················· | ···················· | ····················· | ···················· | ···················· | ···················· |
| H | 8192 | hCoV-19/USA/TX-HMH-1732/2020\|EPI_ISL_542799\|2020-0 | ···················· | ···················· | ····················· | ···················· | ···················· | ···················· |
| H | 8193 | hCoV-19/England/QEUH-96D976/2020\|EPI_ISL_530643\|20 | ···················· | ·A·················· | ····················· | ···················· | ···················· | ···················· |
| H | 8194 | hCoV-19/England/QEUH-96CA29/2020\|EPI_ISL_530644\|20 | ···················· | ·A·················· | ····················· | ···················· | ···················· | ···················· |
| H | 8195 | hCoV-19/Northern Ireland/QEUH-96C92C/2020\|EPI_ISL_ | ···················· | ···················· | ····················· | ···················· | ···················· | ···················· |
| H | 8196 | hCoV-19/Northern Ireland/QEUH-96D3A5/2020\|EPI_ISL_ | ···················· | ······C············· | ····················· | ···················· | ···················· | ···················· |
| H | 8197 | hCoV-19/Northern Ireland/QEUH-96DCAA/2020\|EPI_ISL_ | ···················· | ···················· | ····················· | ···················· | ···················· | ···················· |
| H | 8198 | hCoV-19/Northern Ireland/QEUH-96CF84/2020\|EPI_ISL_ | ···················· | ···················· | ····················· | ···················· | ···················· | ···················· |
| H | 8199 | hCoV-19/Northern Ireland/QEUH-96DA37/2020\|EPI_ISL_ | ···················· | ···················· | ····················· | ···················· | ···················· | ···················· |
| H | 8200 | hCoV-19/Northern Ireland/QEUH-96D062/2020\|EPI_ISL_ | ···················· | ···················· | ····················· | ···················· | ···················· | ···················· |
| H | 8201 | hCoV-19/England/QEUH-96C86B/2020\|EPI_ISL_530656\|20 | ···················· | ···················· | ····················· | ···················· | ···················· | ···················· |
| H | 8202 | hCoV-19/Scotland/QEUH-96C78C/2020\|EPI_ISL_530659\|2 | ···················· | ····A··············· | ····················· | ···················· | ···················· | ···················· |
| H | 8203 | hCoV-19/Northern Ireland/QEUH-96CC23/2020\|EPI_ISL_ | ···················· | ···················· | ····················· | ···················· | ···················· | ···················· |
| H | 8204 | hCoV-19/England/QEUH-96D026/2020\|EPI_ISL_530661\|20 | ···················· | ···················· | ····················· | ···················· | ···················· | ···················· |
| H | 8205 | hCoV-19/Northern Ireland/QEUH-96DB8F/2020\|EPI_ISL_ | ···················· | ···················· | ····················· | ···················· | ···················· | ···················· |
| H | 8206 | hCoV-19/Scotland/QEUH-96121D/2020\|EPI_ISL_531973\|2 | ···················· | ···················· | ····················· | ···················· | ···················· | ···················· |
| H | 8207 | hCoV-19/England/QEUH-9605BB/2020\|EPI_ISL_531974\|20 | ···················· | ···················· | ····················· | ···················· | ···················· | ···················· |
| H | 8208 | hCoV-19/England/QEUH-960287/2020\|EPI_ISL_531975\|20 | ···················· | ···················· | ····················· | ···················· | ···················· | ···················· |
| H | 8209 | hCoV-19/England/QEUH-960418/2020\|EPI_ISL_531976\|20 | ···················· | ···················· | ····················· | ···················· | ···················· | ···················· |
| H | 8210 | hCoV-19/Scotland/QEUH-960F35/2020\|EPI_ISL_531978\|2 | ···················· | ···················· | ····················· | ···················· | ···················· | ···················· |
| H | 8211 | hCoV-19/Scotland/QEUH-960C10/2020\|EPI_ISL_531979\|2 | ···················· | ···················· | ····················· | ···················· | ···················· | ···················· |
| H | 8212 | hCoV-19/England/QEUH-960C98/2020\|EPI_ISL_531980\|20 | ···················· | ···················· | ····················· | ···················· | ···················· | ···················· |
| H | 8213 | hCoV-19/England/QEUH-960D86/2020\|EPI_ISL_531981\|20 | ···················· | ···················· | ····················· | ···················· | ···················· | ···················· |
| H | 8214 | hCoV-19/Scotland/QEUH-9606E5/2020\|EPI_ISL_531982\|2 | ···················· | ···················· | ····················· | ···················· | ···················· | ···················· |
| H | 8215 | hCoV-19/England/QEUH-96105F/2020\|EPI_ISL_531983\|20 | ···················· | ···················· | ····················· | ···················· | ···················· | ···················· |
| H | 8216 | hCoV-19/Scotland/QEUH-960E83/2020\|EPI_ISL_531984\|2 | ···················· | ···················· | ····················· | ···················· | ···················· | ···················· |
| H | 8217 | hCoV-19/Northern Ireland/QEUH-960E65/2020\|EPI_ISL_ | ···················· | ···················· | ····················· | ···················· | ···················· | ···················· |
| H | 8218 | hCoV-19/Northern Ireland/QEUH-9609A0/2020\|EPI_ISL_ | ···················· | ···················· | ····················· | ···················· | ···················· | ···················· |
| H | 8219 | hCoV-19/England/QEUH-9602A5/2020\|EPI_ISL_531988\|20 | ···················· | ······A············· | ····················· | ···················· | ···················· | ···················· |
| H | 8220 | hCoV-19/England/QEUH-9607F1/2020\|EPI_ISL_531989\|20 | ···················· | ···················· | ····················· | ···················· | ···················· | ···················· |
| H | 8221 | hCoV-19/Scotland/QEUH-960560/2020\|EPI_ISL_531990\|2 | ···················· | ···················· | ····················· | ···················· | ···················· | ···················· |
| H | 8222 | hCoV-19/England/QEUH-9603C0/2020\|EPI_ISL_531991\|20 | ···················· | ···················· | ····················· | ···················· | ···················· | ···················· |
| H | 8223 | hCoV-19/England/QEUH-960D77/2020\|EPI_ISL_531993\|20 | ···················· | ·A·················· | ····················· | ···················· | ···················· | ···A················ |
| H | 8224 | hCoV-19/England/QEUH-960472/2020\|EPI_ISL_531994\|20 | ···················· | ···················· | ····················· | ···················· | ···················· | ···················· |
| H | 8225 | hCoV-19/England/QEUH-9608B2/2020\|EPI_ISL_531996\|20 | ···················· | ···················· | ····················· | ···················· | ···················· | ···················· |
| H | 8226 | hCoV-19/South Africa/KRISP-K002504/2020\|EPI_ISL_53 | ···················· | ···················· | ····················· | ···················· | ···················· | ···················· |
| H | 8227 | hCoV-19/South Africa/KRISP-K002520/2020\|EPI_ISL_53 | ···················· | ···················· | ····················· | ···················· | ···················· | ···················· |
| H | 8228 | hCoV-19/South Africa/KRISP-K002526/2020\|EPI_ISL_53 | ···················· | ···················· | ····················· | ···················· | ···················· | ···················· |
| H | 8229 | hCoV-19/South Africa/KRISP-K002532/2020\|EPI_ISL_53 | ···················· | ····A··············· | ····················· | ···················· | ···················· | ···················· |
| H | 8230 | hCoV-19/South Africa/KRISP-K002533/2020\|EPI_ISL_53 | ···················· | ···················· | ····················· | ···················· | ···················· | ···················· |
| H | 8231 | hCoV-19/South Africa/KRISP-K002535/2020\|EPI_ISL_53 | ···················· | ···················· | ····················· | ···················· | ···················· | ···················· |
| H | 8232 | hCoV-19/South Africa/KRISP-K002537/2020\|EPI_ISL_53 | ···················· | ···················· | ····················· | ···················· | ···················· | ···················· |
| H | 8233 | hCoV-19/South Africa/KRISP-K002542/2020\|EPI_ISL_53 | ···················· | ···················· | ····················· | ···················· | ···················· | ···················· |
| H | 8234 | hCoV-19/South Africa/KRISP-K002543/2020\|EPI_ISL_53 | ···················· | ···················· | ····················· | ···················· | ···················· | ···················· |
| H | 8235 | hCoV-19/South Africa/KRISP-K002544/2020\|EPI_ISL_53 | ···················· | ···················· | ····················· | ···················· | ···················· | ···················· |
| H | 8236 | hCoV-19/South Africa/KRISP-K002545/2020\|EPI_ISL_53 | ···················· | ···················· | ····················· | ···················· | ···················· | ···················· |
| H | 8237 | hCoV-19/South Africa/KRISP-K002546/2020\|EPI_ISL_53 | ···················· | ···················· | ····················· | ···················· | ···················· | ···················· |
| H | 8238 | hCoV-19/South Africa/KRISP-K002547/2020\|EPI_ISL_53 | ···················· | ···················· | ····················· | ···················· | ···················· | ···················· |
| H | 8239 | hCoV-19/South Africa/KRISP-K002550/2020\|EPI_ISL_53 | ···················· | ···················· | ····················· | ···················· | ···················· | ···················· |
| H | 8240 | hCoV-19/South Africa/KRISP-K002552/2020\|EPI_ISL_53 | ···················· | ···················· | ····················· | ···················· | ···················· | ···················· |
| H | 8241 | hCoV-19/South Africa/KRISP-K002553/2020\|EPI_ISL_53 | ···················· | ···················· | ····················· | ···················· | ···················· | ···················· |
| H | 8242 | hCoV-19/South Africa/KRISP-K002555/2020\|EPI_ISL_53 | ···················· | ···················· | ····················· | ···················· | ···················· | ···················· |
| H | 8243 | hCoV-19/South Africa/KRISP-K002556/2020\|EPI_ISL_53 | ···················· | ···················· | ····················· | ···················· | ···················· | ···················· |
| H | 8244 | hCoV-19/South Africa/KRISP-K002558/2020\|EPI_ISL_53 | ···················· | ···················· | ····················· | ···················· | ···················· | ···················· |
| H | 8245 | hCoV-19/South Africa/KRISP-K002559/2020\|EPI_ISL_53 | ···················· | ···················· | ····················· | ···················· | ···················· | ···················· |
| H | 8246 | hCoV-19/South Africa/KRISP-K002560/2020\|EPI_ISL_53 | ···················· | ···················· | ····················· | ···················· | ···················· | ···················· |
| H | 8247 | hCoV-19/South Africa/KRISP-K002562/2020\|EPI_ISL_53 | ···················· | ···················· | ····················· | ···················· | ···················· | ···················· |

| | | | | | | | | |
|---|---|---|---|---|---|---|---|---|
| H | 8248 | hCoV-19/South Africa/KRISP-K002563/2020\|EPI_ISL_53 | .................... | .................... | .................... | .................... | .................... | .................... |
| H | 8249 | hCoV-19/South Africa/KRISP-K002566/2020\|EPI_ISL_53 | .................... | .................... | .................... | .................... | .................... | .................... |
| H | 8250 | hCoV-19/South Africa/KRISP-K002567/2020\|EPI_ISL_53 | .................... | .................... | .................... | .................... | .................... | .................... |
| H | 8251 | hCoV-19/South Africa/KRISP-K002568/2020\|EPI_ISL_53 | .......C............ | .................... | .................... | .................... | .................... | .................... |
| H | 8252 | hCoV-19/South Africa/KRISP-K002569/2020\|EPI_ISL_53 | .......C............ | .................... | .................... | .................... | .................... | .................... |
| H | 8253 | hCoV-19/South Africa/KRISP-K002570/2020\|EPI_ISL_53 | .................... | .................... | .................... | .................... | .................... | .................... |
| H | 8254 | hCoV-19/South Africa/KRISP-K002572/2020\|EPI_ISL_53 | .................... | .................... | .................... | .................... | .................... | .................... |
| H | 8255 | hCoV-19/South Africa/KRISP-K002573/2020\|EPI_ISL_53 | .......C............ | .................... | .................... | .................... | .................... | .................... |
| H | 8256 | hCoV-19/South Africa/KRISP-K002574/2020\|EPI_ISL_53 | .................... | .................... | .................... | .................... | .................... | .................... |
| H | 8257 | hCoV-19/South Africa/KRISP-K002576/2020\|EPI_ISL_53 | .................... | .................... | .................... | .................... | .................... | .................... |
| H | 8258 | hCoV-19/South Africa/KRISP-K002577/2020\|EPI_ISL_53 | .................... | .................... | .................... | .................... | .................... | .................... |
| H | 8259 | hCoV-19/South Africa/KRISP-K002578/2020\|EPI_ISL_53 | .................... | .................... | .................... | .................... | .................... | .................... |
| H | 8260 | hCoV-19/South Africa/KRISP-K002579/2020\|EPI_ISL_53 | .................... | ......C............. | .................... | .................... | .................... | .................... |
| H | 8261 | hCoV-19/South Africa/KRISP-K002580/2020\|EPI_ISL_53 | .................... | .................... | .................... | .................... | .................... | .................... |
| H | 8262 | hCoV-19/South Africa/KRISP-K002581/2020\|EPI_ISL_53 | .................... | .................... | .................... | .................... | .................... | .................... |
| H | 8263 | hCoV-19/South Africa/KRISP-K002582/2020\|EPI_ISL_53 | .................... | .................... | .................... | .................... | .................... | .................... |
| H | 8264 | hCoV-19/South Africa/KRISP-K002583/2020\|EPI_ISL_53 | .................... | .................... | .................... | .................... | .................... | .................... |
| H | 8265 | hCoV-19/South Africa/KRISP-K002584/2020\|EPI_ISL_53 | .................... | .................... | .................... | .................... | .................... | .................... |
| H | 8266 | hCoV-19/South Africa/KRISP-K002585/2020\|EPI_ISL_53 | .................... | .................... | .................... | .................... | .................... | .................... |
| H | 8267 | hCoV-19/South Africa/KRISP-K002586/2020\|EPI_ISL_53 | .................... | .................... | .................... | .................... | .................... | .................... |
| H | 8268 | hCoV-19/South Africa/KRISP-K002587/2020\|EPI_ISL_53 | .................... | .................... | .................... | .................... | .................... | ...............A.... |
| H | 8269 | hCoV-19/South Africa/KRISP-K002589/2020\|EPI_ISL_53 | .................... | .................... | .................... | .................... | .................... | .................... |
| H | 8270 | hCoV-19/South Africa/KRISP-K002590/2020\|EPI_ISL_53 | .................... | .................... | .................... | .................... | .................... | .................... |
| H | 8271 | hCoV-19/South Africa/KRISP-K002591/2020\|EPI_ISL_53 | .................... | .................... | .................... | .................... | .................... | .................... |
| H | 8272 | hCoV-19/South Africa/KRISP-K002592/2020\|EPI_ISL_53 | .................... | .................... | .................... | .................... | .................... | .................... |
| H | 8273 | hCoV-19/South Africa/KRISP-K002593/2020\|EPI_ISL_53 | .................... | .................... | .................... | .................... | .................... | .................... |
| H | 8274 | hCoV-19/South Africa/KRISP-K002594/2020\|EPI_ISL_53 | .................... | .................... | .................... | .................... | .................... | .................... |
| H | 8275 | hCoV-19/South Africa/KRISP-K002595/2020\|EPI_ISL_53 | .................... | .................... | .................... | .................... | .................... | .................... |
| H | 8276 | hCoV-19/South Africa/KRISP-K002596/2020\|EPI_ISL_53 | .................... | .................... | .................... | .................... | .................... | .................... |
| H | 8277 | hCoV-19/South Africa/KRISP-K002597/2020\|EPI_ISL_53 | .................... | .................... | .................... | .................... | .................... | .................... |
| H | 8278 | hCoV-19/South Africa/KRISP-K002598/2020\|EPI_ISL_53 | .................... | .................... | .................... | .................... | .................... | .................... |
| H | 8279 | hCoV-19/South Africa/KRISP-K002600/2020\|EPI_ISL_53 | .................... | .................... | .................... | .................... | .................... | .................... |
| H | 8280 | hCoV-19/South Africa/KRISP-K002601/2020\|EPI_ISL_53 | .................... | .................... | .................... | .................... | .................... | .................... |
| H | 8281 | hCoV-19/South Africa/KRISP-K002602/2020\|EPI_ISL_53 | .................... | .................... | .................... | .................... | .................... | .................... |
| H | 8282 | hCoV-19/South Africa/KRISP-K002603/2020\|EPI_ISL_53 | .................... | .................... | .................... | .................... | .................... | .................... |
| H | 8283 | hCoV-19/South Africa/KRISP-K002604/2020\|EPI_ISL_53 | .................... | .................... | .................... | .................... | .................... | .................... |
| H | 8284 | hCoV-19/South Africa/KRISP-K002605/2020\|EPI_ISL_53 | .................... | .................... | .................... | .................... | .................... | .................... |
| H | 8285 | hCoV-19/South Africa/KRISP-K002606/2020\|EPI_ISL_53 | .................... | .................... | .................... | .................... | .................... | .................... |
| H | 8286 | hCoV-19/South Africa/KRISP-K002607/2020\|EPI_ISL_53 | .................... | .................... | .................... | .................... | .................... | .................... |
| H | 8287 | hCoV-19/South Africa/KRISP-K002608/2020\|EPI_ISL_53 | .................... | .................... | .................... | .................... | .................... | .................... |
| H | 8288 | hCoV-19/South Africa/KRISP-K002609/2020\|EPI_ISL_53 | .................... | .................... | .................... | .................... | .................... | .................... |
| H | 8289 | hCoV-19/South Africa/KRISP-K002611/2020\|EPI_ISL_53 | .................... | .................... | .................... | .................... | .................... | .................... |
| H | 8290 | hCoV-19/South Africa/KRISP-K002612/2020\|EPI_ISL_53 | .................... | .................... | .................... | .................... | .................... | .................... |
| H | 8291 | hCoV-19/South Africa/KRISP-K002613/2020\|EPI_ISL_53 | .................... | .................... | .................... | .................... | .................... | .................... |
| H | 8293 | hCoV-19/South Africa/KRISP-K002615/2020\|EPI_ISL_53 | .................... | .................... | .................... | .................... | .................... | .................... |
| H | 8294 | hCoV-19/South Africa/KRISP-K002616/2020\|EPI_ISL_53 | .................... | .................... | .................... | .................... | .................... | .................... |
| H | 8295 | hCoV-19/South Africa/KRISP-K002617/2020\|EPI_ISL_53 | .................... | .................... | .................... | .................... | .................... | .................... |
| H | 8296 | hCoV-19/South Africa/KRISP-K002619/2020\|EPI_ISL_53 | .................... | .................... | .................... | .................... | .................... | .................... |
| H | 8297 | hCoV-19/South Africa/KRISP-K002621/2020\|EPI_ISL_53 | .................... | .................... | .................... | .................... | .................... | .................... |
| H | 8298 | hCoV-19/South Africa/KRISP-K002622/2020\|EPI_ISL_53 | .................... | .................... | .................... | .................... | .................... | .................... |
| H | 8299 | hCoV-19/South Africa/KRISP-K002624/2020\|EPI_ISL_53 | .................... | .................... | .................... | .................... | .................... | .................... |
| H | 8300 | hCoV-19/South Africa/KRISP-K002625/2020\|EPI_ISL_53 | .................... | .................... | .................... | .................... | .................... | .................... |
| H | 8301 | hCoV-19/South Africa/KRISP-K002626/2020\|EPI_ISL_53 | .................... | .................... | .................... | .................... | .................... | .................... |
| H | 8302 | hCoV-19/South Africa/KRISP-K002627/2020\|EPI_ISL_53 | .................... | .................... | .................... | .................... | .................... | .................... |
| H | 8303 | hCoV-19/South Africa/KRISP-K002628/2020\|EPI_ISL_53 | .................... | .................... | .................... | .................... | .................... | .................... |
| H | 8304 | hCoV-19/South Africa/KRISP-K002629/2020\|EPI_ISL_53 | .................... | .................... | .................... | .................... | .................... | .................... |
| H | 8305 | hCoV-19/USA/WI-UW-1329/2020\|EPI_ISL_536700\|2020-08 | .................... | .................... | .................... | .................... | .................... | .................... |
| H | 8306 | hCoV-19/USA/WI-UW-1330/2020\|EPI_ISL_536701\|2020-08 | .................... | ..C................. | .................... | .................... | .................... | .................... |
| H | 8307 | hCoV-19/USA/NY-UW-1331/2020\|EPI_ISL_536702\|2020-08 | .................... | .................... | .................... | .................... | .................... | .................... |
| H | 8308 | hCoV-19/USA/WI-UW-1332/2020\|EPI_ISL_536703\|2020-08 | .................... | .................... | .................... | .................... | .................... | .................... |
| H | 8309 | hCoV-19/USA/WI-UW-1333/2020\|EPI_ISL_536704\|2020-08 | .................... | .................... | .................... | .................... | .................... | .................... |
| H | 8310 | hCoV-19/USA/WI-UW-1334/2020\|EPI_ISL_536705\|2020-08 | .................... | .............A...... | .................... | .................... | .................... | .................... |
| H | 8311 | hCoV-19/USA/WI-UW-1335/2020\|EPI_ISL_536706\|2020-08 | .................... | .................... | .................... | .................... | .................... | .................... |
| H | 8312 | hCoV-19/USA/WI-UW-1336/2020\|EPI_ISL_536707\|2020-09 | .................... | .................... | .................... | .................... | .................... | .................... |
| H | 8313 | hCoV-19/USA/WI-UW-1337/2020\|EPI_ISL_536708\|2020-09 | .................... | .................... | .................... | .................... | .................... | .................... |
| H | 8314 | hCoV-19/USA/WI-UW-1338/2020\|EPI_ISL_536709\|2020-09 | .................... | .................... | .................... | .................... | .................... | .................... |
| H | 8316 | hCoV-19/USA/WI-UW-1340/2020\|EPI_ISL_536711\|2020-09 | .................... | .................... | .................... | .................... | .................... | .................... |
| H | 8317 | hCoV-19/USA/WI-UW-1341/2020\|EPI_ISL_536712\|2020-09 | .................... | .................... | .................... | .................... | .................... | .................... |
| H | 8319 | hCoV-19/USA/NY-UW-1343/2020\|EPI_ISL_536714\|2020-09 | .................... | .................... | .................... | .................... | .................... | .................... |
| H | 8320 | hCoV-19/USA/WI-UW-1344/2020\|EPI_ISL_536715\|2020-09 | .................... | .................... | .................... | .................... | .................... | .................... |
| H | 8321 | hCoV-19/USA/WI-UW-1346/2020\|EPI_ISL_536717\|2020-09 | .................... | .................... | .................... | .................... | .................... | .................... |
| H | 8322 | hCoV-19/USA/WI-UW-1347/2020\|EPI_ISL_536718\|2020-09 | .................... | .................... | .................... | .................... | .................... | .................... |
| H | 8323 | hCoV-19/USA/WI-UW-1348/2020\|EPI_ISL_536719\|2020-09 | .................... | .................... | .................... | .................... | .................... | .................... |
| H | 8324 | hCoV-19/USA/WI-UW-1349/2020\|EPI_ISL_536720\|2020-09 | .................... | .................... | .................... | .................... | .................... | .................... |
| H | 8325 | hCoV-19/USA/WI-UW-1350/2020\|EPI_ISL_536721\|2020-09 | .................... | .................... | .................... | .................... | .................... | .................... |
| H | 8326 | hCoV-19/USA/NY-UW-1351/2020\|EPI_ISL_536722\|2020-09 | .................... | .................... | .................... | .................... | .................... | .................... |
| H | 8327 | hCoV-19/USA/WI-UW-1352/2020\|EPI_ISL_536723\|2020-09 | .................... | .................... | .................... | .................... | .................... | .................... |
| H | 8328 | hCoV-19/USA/WI-UW-1353/2020\|EPI_ISL_536724\|2020-09 | .................... | .................... | .................... | .................... | .................... | .................... |
| H | 8329 | hCoV-19/USA/WI-UW-1354/2020\|EPI_ISL_536725\|2020-09 | .................... | .................... | .................... | .................... | .................... | .................... |
| H | 8330 | hCoV-19/USA/WI-UW-1356/2020\|EPI_ISL_536727\|2020-09 | .................... | .................... | .................... | .................... | .................... | .................... |

| | | | | | | | | | |
|---|---|---|---|---|---|---|---|---|---|
| H | 8331 | hCoV-19/USA/WI-UW-1357/2020\|EPI_ISL_536728\|2020-09 | ···················· | ···················· | ···················· | ···················· | ···················· | ···················· | |
| H | 8332 | hCoV-19/USA/WI-UW-1358/2020\|EPI_ISL_536729\|2020-09 | ···················· | ···················· | ···················· | ···················· | ···················· | ···················· | |
| H | 8333 | hCoV-19/USA/WI-UW-1359/2020\|EPI_ISL_536730\|2020-09 | ···················· | ···················· | ···················· | ···················· | ···················· | ···················· | |
| H | 8334 | hCoV-19/USA/WI-UW-1360/2020\|EPI_ISL_536731\|2020-09 | ···················· | ···················· | ···················· | ···················· | ···················· | ···················· | |
| H | 8335 | hCoV-19/USA/WI-UW-1361/2020\|EPI_ISL_536732\|2020-09 | ···················· | ···················· | ···················· | ···················· | ···················· | ···················· | |
| H | 8336 | hCoV-19/USA/WI-UW-1362/2020\|EPI_ISL_536733\|2020-09 | ···················· | ···················· | ···················· | ···················· | ···················· | ···················· | |
| H | 8337 | hCoV-19/USA/WI-UW-1363/2020\|EPI_ISL_536734\|2020-09 | ···················· | ·A·················· | ···················· | ···················· | ···················· | ···················· | |
| H | 8338 | hCoV-19/USA/WI-UW-1364/2020\|EPI_ISL_536735\|2020-09 | ···················· | ···················· | ···················· | ···················· | ···················· | ···················· | |
| H | 8339 | hCoV-19/USA/NJ-UW-1365/2020\|EPI_ISL_536736\|2020-09 | ···················· | ···················· | ···················· | ···················· | ···················· | ···················· | |
| H | 8340 | hCoV-19/USA/WI-UW-1366/2020\|EPI_ISL_536737\|2020-09 | ···················· | ···················· | ···················· | ···················· | ···················· | ···················· | |
| H | 8341 | hCoV-19/USA/WI-UW-1367/2020\|EPI_ISL_536738\|2020-09 | ···················· | ···················· | ···················· | ···················· | ···················· | ···················· | |
| H | 8342 | hCoV-19/USA/WI-UW-1368/2020\|EPI_ISL_536739\|2020-09 | ···················· | ···················· | ···················· | ···················· | ···················· | ···················· | |
| H | 8343 | hCoV-19/USA/WI-UW-1369/2020\|EPI_ISL_536740\|2020-09 | ···················· | ···················· | ···················· | ···················· | ···················· | ···················· | |
| H | 8344 | hCoV-19/USA/WI-UW-1370/2020\|EPI_ISL_536741\|2020-09 | ···················· | ···················· | ···················· | ···················· | ···················· | ···················· | |
| H | 8345 | hCoV-19/USA/WI-UW-1371/2020\|EPI_ISL_536742\|2020-09 | ···················· | ···················· | ···················· | ···················· | ···················· | ···················· | |
| H | 8346 | hCoV-19/USA/WI-UW-1372/2020\|EPI_ISL_536743\|2020-09 | ···················· | ···················· | ···················· | ···················· | ···················· | ···················· | |
| H | 8347 | hCoV-19/USA/WI-UW-1373/2020\|EPI_ISL_536744\|2020-09 | ···················· | ···················· | ···················· | ···················· | ···················· | ···················· | |
| H | 8348 | hCoV-19/USA/WI-UW-1374/2020\|EPI_ISL_536745\|2020-09 | ···················· | ···················· | ···················· | ···················· | ···················· | ···················· | |
| H | 8349 | hCoV-19/USA/WI-UW-1375/2020\|EPI_ISL_536746\|2020-09 | ···················· | ···················· | ···················· | ···················· | ···················· | ···················· | |
| H | 8350 | hCoV-19/USA/WI-UW-1376/2020\|EPI_ISL_536747\|2020-09 | ···················· | ···················· | ···················· | ···················· | ···················· | ···················· | |
| H | 8351 | hCoV-19/USA/WI-UW-1379/2020\|EPI_ISL_536750\|2020-09 | ···················· | ···················· | ···················· | ···················· | ···················· | ···················· | |
| H | 8352 | hCoV-19/USA/NY-UW-1381/2020\|EPI_ISL_536752\|2020-09 | ···················· | ···················· | ···················· | ···················· | ···················· | ···················· | |
| H | 8353 | hCoV-19/USA/WI-UW-1382/2020\|EPI_ISL_536753\|2020-09 | ···················· | ···················· | ···················· | ···················· | ···················· | ···················· | |
| H | 8355 | hCoV-19/USA/WI-UW-1384/2020\|EPI_ISL_536755\|2020-09 | ···················· | ···················· | ···················· | ···················· | ···················· | ···················· | |
| H | 8356 | hCoV-19/USA/WI-UW-1385/2020\|EPI_ISL_536756\|2020-09 | ···················· | ···················· | ···················· | ···················· | ···················· | ···················· | |
| H | 8357 | hCoV-19/USA/WI-UW-1386/2020\|EPI_ISL_536757\|2020-09 | ···················· | ···················· | ···················· | ···················· | ···················· | ···················· | |
| H | 8358 | hCoV-19/USA/FL-UW-1387/2020\|EPI_ISL_536758\|2020-09 | ···················· | ···················· | ···················· | ···················· | ···················· | ···················· | |
| H | 8359 | hCoV-19/USA/WI-UW-1388/2020\|EPI_ISL_536759\|2020-09 | ···················· | ···················· | ···················· | ···················· | ···················· | ···················· | |
| H | 8360 | hCoV-19/USA/NY-UW-1389/2020\|EPI_ISL_536760\|2020-09 | ···················· | ···················· | ···················· | ···················· | ···················· | ···················· | |
| H | 8361 | hCoV-19/USA/WI-UW-1390/2020\|EPI_ISL_536761\|2020-09 | ···················· | ···················· | ···················· | ···················· | ···················· | ···················· | |
| H | 8362 | hCoV-19/USA/WI-UW-1391/2020\|EPI_ISL_536762\|2020-09 | ···················· | ···················· | ···················· | ···················· | ···················· | ···················· | |
| H | 8363 | hCoV-19/USA/MO-UW-1392/2020\|EPI_ISL_536763\|2020-09 | ···················· | ···················· | ···················· | ···················· | ···················· | ···················· | |
| H | 8364 | hCoV-19/USA/WI-UW-1393/2020\|EPI_ISL_536764\|2020-09 | ···················· | ···················· | ···················· | ···················· | ···················· | ···················· | |
| H | 8365 | hCoV-19/USA/WI-UW-1394/2020\|EPI_ISL_536765\|2020-09 | ···················· | ···················· | ···················· | ···················· | ···················· | ···················· | |
| H | 8366 | hCoV-19/USA/WI-UW-1395/2020\|EPI_ISL_536766\|2020-09 | ···················· | ···················· | ···················· | ···················· | ···················· | ···················· | |
| H | 8367 | hCoV-19/USA/WI-UW-1396/2020\|EPI_ISL_536767\|2020-09 | ···················· | ···················· | ···················· | ···················· | ···················· | ···················· | |
| H | 8368 | hCoV-19/USA/WI-UW-1397/2020\|EPI_ISL_536768\|2020-09 | ···················· | ···················· | ···················· | ···················· | ···················· | ···················· | |
| H | 8369 | hCoV-19/USA/WI-UW-1398/2020\|EPI_ISL_536769\|2020-09 | ···················· | ···················· | ···················· | ···················· | ···················· | ···················· | |
| H | 8370 | hCoV-19/USA/WI-UW-1401/2020\|EPI_ISL_536772\|2020-09 | ···················· | ···················· | ···················· | ···················· | ···················· | ···················· | |
| H | 8371 | hCoV-19/USA/NY-UW-1403/2020\|EPI_ISL_536773\|2020-09 | ···················· | ···················· | ···················· | ···················· | ···················· | ···················· | |
| H | 8372 | hCoV-19/USA/WI-UW-1404/2020\|EPI_ISL_536774\|2020-09 | ···················· | ···················· | ···················· | ···················· | ···················· | ···················· | |
| H | 8373 | hCoV-19/USA/WI-UW-1405/2020\|EPI_ISL_536775\|2020-09 | ···················· | ···················· | ···················· | ···················· | ···················· | ···················· | |
| H | 8374 | hCoV-19/USA/WI-UW-1406/2020\|EPI_ISL_536776\|2020-09 | ···················· | ···················· | ···················· | ···················· | ···················· | ···················· | |
| H | 8375 | hCoV-19/USA/WI-UW-1407/2020\|EPI_ISL_536777\|2020-09 | ···················· | ···················· | ···················· | ···················· | ···················· | ···················· | |
| H | 8376 | hCoV-19/USA/WI-UW-1408/2020\|EPI_ISL_536778\|2020-09 | ···················· | ···················· | ···················· | ···················· | ···················· | ···················· | |
| H | 8377 | hCoV-19/USA/WI-UW-1409/2020\|EPI_ISL_536779\|2020-09 | ···················· | ···················· | ···················· | ···················· | ···················· | ···················· | |
| H | 8378 | hCoV-19/USA/WI-UW-1410/2020\|EPI_ISL_536780\|2020-09 | ···················· | ···················· | ···················· | ···················· | ···················· | ···················· | |
| H | 8379 | hCoV-19/USA/WI-UW-1411/2020\|EPI_ISL_536781\|2020-09 | ···················· | ···················· | ···················· | ···················· | ···················· | ···················· | |
| H | 8380 | hCoV-19/USA/WI-UW-1412/2020\|EPI_ISL_536782\|2020-09 | ···················· | ···················· | ···················· | ···················· | ···················· | ···················· | |
| H | 8381 | hCoV-19/USA/WI-UW-1413/2020\|EPI_ISL_536783\|2020-09 | ···················· | ···················· | ···················· | ···················· | ···················· | ···················· | |
| H | 8382 | hCoV-19/USA/WI-UW-1414/2020\|EPI_ISL_536784\|2020-09 | ···················· | ···················· | ···················· | ···················· | ···················· | ···················· | |
| H | 8383 | hCoV-19/USA/WI-UW-1415/2020\|EPI_ISL_536785\|2020-08 | ···················· | ···················· | ···················· | ···················· | ···················· | ···················· | |
| H | 8384 | hCoV-19/USA/WI-UW-1416/2020\|EPI_ISL_536786\|2020-09 | ···················· | ···················· | ···················· | ···················· | ···················· | ···················· | |
| H | 8385 | hCoV-19/New Zealand/20VR1765/2020\|EPI_ISL_536789\|2 | ···················· | ···················· | ···················· | ···················· | ···················· | ···················· | |
| H | 8386 | hCoV-19/Hong Kong/MV2007-00828/2020\|EPI_ISL_536793 | ···················· | ···················· | ···················· | ···················· | ···················· | ···················· | |
| H | 8387 | hCoV-19/USA/FL-BPHL-1319/2020\|EPI_ISL_541257\|2020- | ···················· | ···················· | ···················· | ···················· | ···················· | ···················· | |
| H | 8388 | hCoV-19/USA/FL-BPHL-1321/2020\|EPI_ISL_541259\|2020- | ···················· | ···················· | ···················· | ···················· | ···················· | ···················· | |
| H | 8389 | hCoV-19/USA/FL-BPHL-1323/2020\|EPI_ISL_541261\|2020- | ···················· | ···················· | ···················· | ···················· | ···················· | ···················· | |
| H | 8392 | hCoV-19/USA/FL-BPHL-1326/2020\|EPI_ISL_541264\|2020- | ···················· | ···················· | ···················· | ···················· | ···················· | ···················· | |
| H | 8393 | hCoV-19/USA/FL-BPHL-1327/2020\|EPI_ISL_541265\|2020- | ···················· | ···················· | ···················· | ···················· | ···················· | ···················· | |
| H | 8395 | hCoV-19/USA/FL-BPHL-1329/2020\|EPI_ISL_541267\|2020- | ···················· | ···················· | ···················· | ···················· | ···················· | ············NNN····· | 1 |
| H | 8397 | hCoV-19/USA/FL-BPHL-1331/2020\|EPI_ISL_541269\|2020- | ···················· | ···················· | ···················· | ···················· | ···················· | ···················· | |
| H | 8399 | hCoV-19/USA/FL-BPHL-1334/2020\|EPI_ISL_541272\|2020- | ···················· | ·A·················· | ···················· | ···················· | ···················· | ···················· | |
| H | 8401 | hCoV-19/USA/FL-BPHL-1340/2020\|EPI_ISL_541278\|2020- | ···················· | ···················· | ···················· | ···················· | ···················· | ···················· | |
| H | 8402 | hCoV-19/USA/FL-BPHL-1341/2020\|EPI_ISL_541279\|2020- | ···················· | ···················· | ···················· | ···················· | ···················· | ···················· | |
| H | 8403 | hCoV-19/USA/FL-BPHL-1343/2020\|EPI_ISL_541281\|2020- | ···················· | ···················· | ···················· | ···················· | ···················· | ···················· | |
| H | 8404 | hCoV-19/USA/FL-BPHL-1344/2020\|EPI_ISL_541282\|2020- | ···················· | ···················· | ···················· | ···················· | ···················· | ···················· | |
| H | 8405 | hCoV-19/USA/FL-BPHL-1347/2020\|EPI_ISL_541285\|2020- | ···················· | ···················· | ···················· | ···················· | ···················· | ···················· | |
| H | 8406 | hCoV-19/USA/FL-BPHL-1348/2020\|EPI_ISL_541286\|2020- | ···················· | ···················· | ···················· | ···················· | ···················· | ···················· | |
| H | 8407 | hCoV-19/USA/FL-BPHL-1349/2020\|EPI_ISL_541287\|2020- | ···················· | ···················· | ···················· | ···················· | ···················· | ···················· | |
| H | 8408 | hCoV-19/USA/FL-BPHL-1351/2020\|EPI_ISL_541289\|2020- | ···················· | ···················· | ···················· | ···················· | ···················· | ···················· | |
| H | 8409 | hCoV-19/USA/FL-BPHL-1353/2020\|EPI_ISL_541291\|2020- | ···················· | ···················· | ···················· | ···················· | ···················· | ···················· | |
| H | 8410 | hCoV-19/USA/FL-BPHL-1354/2020\|EPI_ISL_541292\|2020- | ···················· | ···················· | ···················· | ···················· | ···················· | ···················· | |
| H | 8417 | hCoV-19/USA/FL-BPHL-1366/2020\|EPI_ISL_541303\|2020- | ···················· | ···················· | ···················· | ···················· | ···················· | ···················· | |
| H | 8418 | hCoV-19/USA/FL-BPHL-1372/2020\|EPI_ISL_541309\|2020- | ···········T········ | ···················· | ···················· | ···················· | ···················· | ···················· | |
| H | 8419 | hCoV-19/USA/TX-HMH-1218/2020\|EPI_ISL_542588\|2020-0 | ···················· | ···················· | ···················· | ···················· | ···················· | ···················· | |
| H | 8420 | hCoV-19/USA/TX-HMH-1220/2020\|EPI_ISL_542589\|2020-0 | ···················· | ···················· | ···················· | ···················· | ···················· | ···················· | |
| H | 8421 | hCoV-19/USA/TX-HMH-1221/2020\|EPI_ISL_542590\|2020-0 | ···················· | ···················· | ···················· | ···················· | ···················· | ···················· | |
| H | 8422 | hCoV-19/USA/TX-HMH-1222/2020\|EPI_ISL_542591\|2020-0 | ···················· | ···················· | ···················· | ···················· | ···················· | ···················· | |
| H | 8423 | hCoV-19/USA/TX-HMH-1225/2020\|EPI_ISL_542592\|2020-0 | ···················· | ···················· | ···················· | ···················· | ···················· | ···················· | |

| | | | | | | | | |
|---|---|---|---|---|---|---|---|---|
| H | 8424 | hCoV-19/USA/TX-HMH-1226/2020\|EPI_ISL_542593\|2020-0 | ···················· | ···················· | ···················· | ···················· | ···················· | ···················· |
| H | 8425 | hCoV-19/USA/TX-HMH-1228/2020\|EPI_ISL_542594\|2020-0 | ···················· | ···················· | ···················· | ···················· | ···················· | ···················· |
| H | 8426 | hCoV-19/USA/TX-HMH-1229/2020\|EPI_ISL_542595\|2020-0 | ···················· | ···················· | ···················· | ···················· | ···················· | ···················· |
| H | 8427 | hCoV-19/USA/TX-HMH-1231/2020\|EPI_ISL_542596\|2020-0 | ···················· | ···················· | ···················· | ···················· | ···················· | ···················· |
| H | 8428 | hCoV-19/USA/TX-HMH-1232/2020\|EPI_ISL_542597\|2020-0 | ···················· | ···················· | ···················· | ···················· | ···················· | ···················· |
| H | 8429 | hCoV-19/USA/TX-HMH-1233/2020\|EPI_ISL_542598\|2020-0 | ···················· | ···················· | ···················· | ···················· | ···················· | ···················· |
| H | 8430 | hCoV-19/USA/TX-HMH-1234/2020\|EPI_ISL_542599\|2020-0 | ···················· | ···················· | ···················· | ···················· | ···················· | ···················· |
| H | 8431 | hCoV-19/USA/TX-HMH-1271/2020\|EPI_ISL_542621\|2020-0 | ···················· | ···················· | ···················· | ···················· | ···················· | ···················· |
| H | 8432 | hCoV-19/USA/TX-HMH-1272/2020\|EPI_ISL_542622\|2020-0 | ···················· | ···················· | ···················· | ···················· | ···················· | ···················· |
| H | 8433 | hCoV-19/USA/TX-HMH-1273/2020\|EPI_ISL_542623\|2020-0 | ···················· | ···················· | ···················· | ···················· | ···················· | ···················· |
| H | 8434 | hCoV-19/USA/TX-HMH-1274/2020\|EPI_ISL_542624\|2020-0 | ···················· | ···················· | ···················· | ···················· | ···················· | ···················· |
| H | 8435 | hCoV-19/USA/TX-HMH-1278/2020\|EPI_ISL_542625\|2020-0 | ···················· | ···················· | ···················· | ···················· | ···················· | ···················· |
| H | 8436 | hCoV-19/USA/TX-HMH-1279/2020\|EPI_ISL_542626\|2020-0 | ···················· | ···················· | ···················· | ···················· | ···················· | ···················· |
| H | 8437 | hCoV-19/USA/TX-HMH-1281/2020\|EPI_ISL_542628\|2020-0 | ···················· | ···················· | ···················· | ···················· | ···················· | ···················· |
| H | 8438 | hCoV-19/USA/TX-HMH-1282/2020\|EPI_ISL_542629\|2020-0 | ···················· | ···················· | ···················· | ···················· | ···················· | ···················· |
| H | 8439 | hCoV-19/USA/TX-HMH-1286/2020\|EPI_ISL_542630\|2020-0 | ···················· | ···················· | ···················· | ···················· | ···················· | ···················· |
| H | 8440 | hCoV-19/USA/TX-HMH-1291/2020\|EPI_ISL_542631\|2020-0 | ···················· | ···················· | ···················· | ···················· | ···················· | ···················· |
| H | 8441 | hCoV-19/USA/TX-HMH-1296/2020\|EPI_ISL_542634\|2020-0 | ···················· | ···················· | ···················· | ···················· | ···················· | ···················· |
| H | 8442 | hCoV-19/USA/TX-HMH-1298/2020\|EPI_ISL_542635\|2020-0 | ···················· | ···················· | ···················· | ···················· | ···················· | ···················· |
| H | 8443 | hCoV-19/USA/TX-HMH-1301/2020\|EPI_ISL_542637\|2020-0 | ···················· | ···················· | ···················· | ···················· | ···················· | ···················· |
| H | 8445 | hCoV-19/USA/TX-HMH-1304/2020\|EPI_ISL_542639\|2020-0 | ···················· | ···················· | ···················· | ···················· | ···················· | ···················· |
| H | 8446 | hCoV-19/USA/TX-HMH-1305/2020\|EPI_ISL_542640\|2020-0 | ···················· | ···················· | ···················· | ···················· | ···················· | ···················· |
| H | 8448 | hCoV-19/USA/TX-HMH-1308/2020\|EPI_ISL_542642\|2020-0 | ···················· | ···················· | ···················· | ···················· | ···················· | ···················· |
| H | 8449 | hCoV-19/England/QEUH-96E90C/2020\|EPI_ISL_530601\|20 | ···················· | ·A·················· | ···················· | ···················· | ···················· | ···················· |
| H | 8450 | hCoV-19/England/QEUH-96E6E7/2020\|EPI_ISL_530602\|20 | ···················· | ····A··············· | ···················· | ···················· | ···················· | ···················· |
| H | 8451 | hCoV-19/England/QEUH-96F5CB/2020\|EPI_ISL_530603\|20 | ···················· | ···················· | ···················· | ···················· | ···················· | ···················· |
| H | 8452 | hCoV-19/England/QEUH-96E016/2020\|EPI_ISL_530606\|20 | ···················· | ···················· | ···················· | ···················· | ······T············· | ···················· |
| H | 8453 | hCoV-19/England/QEUH-96E492/2020\|EPI_ISL_530607\|20 | ···················· | ···················· | ···················· | ···················· | ···················· | ···················· |
| H | 8454 | hCoV-19/England/QEUH-96F051/2020\|EPI_ISL_530608\|20 | ···················· | ···················· | ···················· | ···················· | ···················· | ···················· |
| H | 8455 | hCoV-19/Northern Ireland/QEUH-96F033/2020\|EPI_ISL_ | ···················· | ···················· | ···················· | ···················· | ···················· | ···················· |
| H | 8456 | hCoV-19/England/QEUH-96D466/2020\|EPI_ISL_530664\|20 | ···················· | ·A·················· | ···················· | ···················· | ···················· | ···················· |
| H | 8457 | hCoV-19/Northern Ireland/QEUH-96DC31/2020\|EPI_ISL_ | ···················· | ···················· | ···················· | ···················· | ···················· | ···················· |
| H | 8458 | hCoV-19/Northern Ireland/QEUH-96DA55/2020\|EPI_ISL_ | ···················· | ···················· | ···················· | ···················· | ···················· | ···················· |
| H | 8459 | hCoV-19/Northern Ireland/QEUH-96CB53/2020\|EPI_ISL_ | ···················· | ···················· | ···················· | ···················· | ···················· | ···················· |
| H | 8460 | hCoV-19/Northern Ireland/QEUH-96DA28/2020\|EPI_ISL_ | ···················· | ···················· | ···················· | ···················· | ···················· | ···················· |
| H | 8461 | hCoV-19/Northern Ireland/QEUH-96DB34/2020\|EPI_ISL_ | ···················· | ···················· | ···················· | ···················· | ···················· | ···················· |
| H | 8462 | hCoV-19/England/QEUH-96D554/2020\|EPI_ISL_530671\|20 | ···················· | ···················· | ···················· | ···················· | ···················· | ···················· |
| H | 8463 | hCoV-19/England/QEUH-96DAEC/2020\|EPI_ISL_530672\|20 | ···················· | ···················· | ···················· | ···················· | ···················· | ···················· |
| H | 8464 | hCoV-19/Scotland/QEUH-96D4C0/2020\|EPI_ISL_530673\|2 | ···················· | ···················· | ···················· | ···················· | ···················· | ···················· |
| H | 8465 | hCoV-19/Northern Ireland/QEUH-96DCB9/2020\|EPI_ISL_ | ···················· | ···················· | ···················· | ···················· | ···················· | ···················· |
| H | 8466 | hCoV-19/Northern Ireland/QEUH-96CFFD/2020\|EPI_ISL_ | ···················· | ···················· | ···················· | ···················· | ···················· | ···················· |
| H | 8467 | hCoV-19/Northern Ireland/QEUH-96CC32/2020\|EPI_ISL_ | ········T··········· | ···················· | ···················· | ···················· | ···················· | ···················· |
| H | 8468 | hCoV-19/Northern Ireland/QEUH-96DBE9/2020\|EPI_ISL_ | ···················· | ···················· | ···················· | ···················· | ···················· | ···················· |
| H | 8469 | hCoV-19/Northern Ireland/QEUH-96CB9F/2020\|EPI_ISL_ | ···················· | ···················· | ···················· | ···················· | ···················· | ···················· |
| H | 8470 | hCoV-19/Scotland/QEUH-96C8E3/2020\|EPI_ISL_530682\|2 | ···················· | ·A·················· | ···················· | ···················· | ···················· | ···A················ |
| H | 8471 | hCoV-19/Northern Ireland/QEUH-96CAED/2020\|EPI_ISL_ | ···················· | ···················· | ···················· | ···················· | ···················· | ···················· |
| H | 8472 | hCoV-19/England/QEUH-96C7F5/2020\|EPI_ISL_530684\|20 | ···················· | ···················· | ···················· | ···················· | ···················· | ···················· |
| H | 8473 | hCoV-19/Northern Ireland/QEUH-96DD10/2020\|EPI_ISL_ | ···················· | ···················· | ···················· | ···················· | ···················· | ···················· |
| H | 8474 | hCoV-19/England/QEUH-96D27B/2020\|EPI_ISL_530687\|20 | ···················· | ·A·················· | ···················· | ···················· | ···················· | ···················· |
| H | 8475 | hCoV-19/Northern Ireland/QEUH-96CFA2/2020\|EPI_ISL_ | ···················· | ···················· | ···················· | ···················· | ···················· | ···················· |
| H | 8476 | hCoV-19/Northern Ireland/QEUH-96DB16/2020\|EPI_ISL_ | ···················· | ···················· | ···················· | ···················· | ···················· | ···················· |
| H | 8477 | hCoV-19/Northern Ireland/QEUH-96DA91/2020\|EPI_ISL_ | ···················· | ·A·················· | ···················· | ···················· | ···················· | ···················· |
| H | 8478 | hCoV-19/England/QEUH-96D17E/2020\|EPI_ISL_530692\|20 | ···················· | ···················· | ···················· | ···················· | ···················· | ···················· |
| H | 8479 | hCoV-19/Northern Ireland/QEUH-96CE5A/2020\|EPI_ISL_ | ···················· | ···················· | ···················· | ···················· | ···················· | ···················· |
| H | 8480 | hCoV-19/England/QEUH-96C84D/2020\|EPI_ISL_530694\|20 | ···················· | ·A·················· | ···················· | ···················· | ···················· | ···················· |
| H | 8481 | hCoV-19/England/QEUH-96D18D/2020\|EPI_ISL_530696\|20 | ···················· | ·A·················· | ···················· | ···················· | ···················· | ···················· |
| H | 8482 | hCoV-19/England/QEUH-96D26C/2020\|EPI_ISL_530697\|20 | ···················· | ···················· | ···················· | ···················· | ···················· | ···················· |
| H | 8483 | hCoV-19/England/QEUH-96CB26/2020\|EPI_ISL_530698\|20 | ···················· | ···················· | ···················· | ···················· | ···················· | ···················· |
| H | 8484 | hCoV-19/Northern Ireland/QEUH-96CBEA/2020\|EPI_ISL_ | ···················· | ···················· | ···················· | ···················· | ···················· | ···················· |
| H | 8485 | hCoV-19/England/QEUH-95FBEA/2020\|EPI_ISL_531900\|20 | ···················· | ·A·················· | ···················· | ···················· | ···················· | ···················· |
| H | 8486 | hCoV-19/England/QEUH-96022D/2020\|EPI_ISL_531901\|20 | ···················· | ···················· | ···················· | ···················· | ···················· | ···················· |
| H | 8487 | hCoV-19/England/QEUH-96021E/2020\|EPI_ISL_531903\|20 | ···················· | ···················· | ···················· | ···················· | ···················· | ···················· |
| H | 8488 | hCoV-19/Scotland/QEUH-960032/2020\|EPI_ISL_531905\|2 | ···················· | ···················· | ···················· | ···················· | ···················· | ···················· |
| H | 8489 | hCoV-19/England/QEUH-95FDD5/2020\|EPI_ISL_531906\|20 | ···················· | ···················· | ···················· | ···················· | ···················· | ···················· |
| H | 8490 | hCoV-19/England/QEUH-95FB62/2020\|EPI_ISL_531907\|20 | ···················· | ·A·················· | ···················· | ···················· | ···················· | ···················· |
| H | 8491 | hCoV-19/England/QEUH-95FBDB/2020\|EPI_ISL_531908\|20 | ···················· | ···················· | ···················· | ···················· | ···················· | ···················· |
| H | 8492 | hCoV-19/England/QEUH-960199/2020\|EPI_ISL_531909\|20 | ···················· | ···················· | ···················· | ···················· | ···················· | ···················· |
| H | 8493 | hCoV-19/England/QEUH-95FDF3/2020\|EPI_ISL_531910\|20 | ···················· | ···················· | ···················· | ···················· | ···················· | ···················· |
| H | 8494 | hCoV-19/Scotland/QEUH-960023/2020\|EPI_ISL_531912\|2 | ···················· | ···················· | ···················· | ···················· | ···················· | ···················· |
| H | 8495 | hCoV-19/Scotland/QEUH-95FCD8/2020\|EPI_ISL_531913\|2 | ···················· | ···················· | ···················· | ···················· | ···················· | ···················· |
| H | 8496 | hCoV-19/England/QEUH-95FE5A/2020\|EPI_ISL_531916\|20 | ···················· | ···················· | ···················· | ···················· | ···················· | ···················· |
| H | 8497 | hCoV-19/England/QEUH-95FED2/2020\|EPI_ISL_531917\|20 | ···················· | ···················· | ···················· | ···················· | ···················· | ···················· |
| H | 8498 | hCoV-19/Scotland/QEUH-95FDB7/2020\|EPI_ISL_531918\|2 | ···················· | ···················· | ···················· | ···················· | ···················· | ···················· |
| H | 8499 | hCoV-19/Scotland/QEUH-95FF66/2020\|EPI_ISL_531919\|2 | ···················· | ···················· | ···················· | ···················· | ···················· | ···················· |
| H | 8500 | hCoV-19/England/QEUH-95FCBA/2020\|EPI_ISL_531920\|20 | ···················· | ···················· | ···················· | ···················· | ···················· | ···················· |
| H | 8501 | hCoV-19/Northern Ireland/QEUH-95FD20/2020\|EPI_ISL_ | ···················· | ···················· | ···················· | ···················· | ···················· | ···················· |
| H | 8502 | hCoV-19/England/QEUH-96018A/2020\|EPI_ISL_531922\|20 | ···················· | ···················· | ···················· | ···················· | ···················· | ···················· |
| H | 8503 | hCoV-19/England/QEUH-95FFC0/2020\|EPI_ISL_531923\|20 | ···················· | ···················· | ···················· | ···················· | ···················· | ···················· |
| H | 8504 | hCoV-19/Northern Ireland/QEUH-95FEC3/2020\|EPI_ISL_ | ···················· | ···················· | ···················· | ···················· | ···················· | ···················· |
| H | 8505 | hCoV-19/Scotland/QEUH-95FFEE/2020\|EPI_ISL_531925\|2 | ···················· | ···················· | ···················· | ···················· | ···················· | ···················· |

| | | | | | | | | | |
|---|---|---|---|---|---|---|---|---|---|
| H | 8506 | hCoV-19/England/QEUH-95FC6F/2020\|EPI_ISL_531926\|20 | ···················· | ···················· | ···················· | ···················· | ···················· | ···················· | |
| H | 8507 | hCoV-19/England/QEUH-960102/2020\|EPI_ISL_531927\|20 | ···················· | ···················· | ···················· | ···················· | ···················· | ···················· | |
| H | 8508 | hCoV-19/Scotland/QEUH-95FE1E/2020\|EPI_ISL_531929\|2 | ···················· | ···················· | ···················· | ···················· | ···················· | ···················· | |
| H | 8509 | hCoV-19/England/QEUH-960005/2020\|EPI_ISL_531930\|20 | ···················· | ···················· | ···················· | ···················· | ···················· | ···················· | |
| H | 8510 | hCoV-19/England/QEUH-96009C/2020\|EPI_ISL_531931\|20 | ···················· | ···················· | ···················· | ···················· | ···················· | ···················· | |
| H | 8511 | hCoV-19/Scotland/QEUH-960041/2020\|EPI_ISL_531932\|2 | ···················· | ···················· | ···················· | ···················· | ···················· | ···················· | |
| H | 8512 | hCoV-19/Scotland/QEUH-961189/2020\|EPI_ISL_531933\|2 | ···················· | ···················· | ···················· | ···················· | ···················· | ···················· | |
| H | 8513 | hCoV-19/England/QEUH-960849/2020\|EPI_ISL_531934\|20 | ···················· | ···················· | ···················· | ···················· | ···················· | ···················· | |
| H | 8514 | hCoV-19/Scotland/QEUH-960D95/2020\|EPI_ISL_531935\|2 | ···················· | ···················· | ···················· | ···················· | ···················· | ···················· | |
| H | 8515 | hCoV-19/Scotland/QEUH-960779/2020\|EPI_ISL_531936\|2 | ···················· | ···················· | ···················· | ···················· | ···················· | ···················· | |
| H | 8516 | hCoV-19/England/QEUH-960A34/2020\|EPI_ISL_531937\|20 | ···················· | ·A·······A·········· | ···················· | ···················· | ···················· | ···················· | |
| H | 8517 | hCoV-19/England/QEUH-960964/2020\|EPI_ISL_531938\|20 | ···················· | ·A·······A·········· | ···················· | ···················· | ···················· | ···················· | |
| H | 8518 | hCoV-19/England/QEUH-9611E3/2020\|EPI_ISL_531939\|20 | ···················· | ···················· | ···················· | ···················· | ···················· | ···················· | |
| H | 8519 | hCoV-19/England/QEUH-961286/2020\|EPI_ISL_531940\|20 | ···················· | ···················· | ···················· | ···················· | ···················· | ···················· | |
| H | 8520 | hCoV-19/Scotland/QEUH-960C2F/2020\|EPI_ISL_531995\|2 | ···················· | ···················· | ···················· | ···················· | ···················· | ···················· | |
| H | 8521 | hCoV-19/England/QEUH-9606D6/2020\|EPI_ISL_531997\|20 | ···················· | ···················· | ···················· | ···················· | ···················· | ···················· | |
| H | 8522 | hCoV-19/England/QEUH-9601B7/2020\|EPI_ISL_531998\|20 | ···················· | ···················· | ···················· | ···················· | ···················· | ···················· | |
| H | 8523 | hCoV-19/England/QEUH-960A70/2020\|EPI_ISL_531999\|20 | ···················· | ···················· | ···················· | ···················· | ···················· | ···················· | |
| H | 8524 | hCoV-19/Latvia/110/2020\|EPI_ISL_534199\|2020-08-06 | ···················· | ···················· | ···················· | ···················· | ···················· | ···················· | |
| H | 8525 | hCoV-19/England/LEED-2A82B7/2020\|EPI_ISL_539000\|20 | ···················· | ···················· | ···················· | ···················· | ···················· | ···················· | |
| H | 8526 | hCoV-19/England/LEED-2A8396/2020\|EPI_ISL_539001\|20 | ···················· | ···················· | ···················· | ···················· | ···················· | ···················· | |
| H | 8527 | hCoV-19/England/LEED-2A825D/2020\|EPI_ISL_539002\|20 | ···················· | ···················· | ···················· | ···················· | ···················· | ···················· | |
| H | 8528 | hCoV-19/England/LEED-2A826C/2020\|EPI_ISL_539003\|20 | ···················· | ···················· | ···················· | ···················· | ···················· | ···················· | |
| H | 8529 | hCoV-19/England/LEED-2A8378/2020\|EPI_ISL_539004\|20 | ···················· | ···················· | ···················· | ···················· | ···················· | ···················· | |
| H | 8530 | hCoV-19/England/LEED-2A893A/2020\|EPI_ISL_539007\|20 | ···················· | ···A················ | ···················· | ···················· | ···T················ | ···················· | |
| H | 8531 | hCoV-19/England/LEED-2A8CE6/2020\|EPI_ISL_539008\|20 | ···················· | ···················· | ···················· | ···················· | ···················· | ···················· | |
| H | 8532 | hCoV-19/England/LEED-2A9052/2020\|EPI_ISL_539010\|20 | ···················· | ···················· | ···················· | ···················· | ···················· | ···················· | |
| H | 8533 | hCoV-19/England/LEED-2A86F7/2020\|EPI_ISL_539013\|20 | ···················· | ···················· | ···················· | ···················· | ···················· | ···················· | |
| H | 8534 | hCoV-19/England/LEED-2A8BAD/2020\|EPI_ISL_539014\|20 | ···················· | ···A················ | ···················· | ···················· | ···················· | ···················· | |
| H | 8535 | hCoV-19/England/LEED-2A8615/2020\|EPI_ISL_539015\|20 | ···················· | ···················· | ···················· | ···················· | ···················· | ···················· | |
| H | 8536 | hCoV-19/England/LEED-2A91D7/2020\|EPI_ISL_539016\|20 | ···················· | ·····A·············· | ···················· | ···················· | ···················· | ···················· | |
| H | 8537 | hCoV-19/England/LEED-2A87C7/2020\|EPI_ISL_539017\|20 | ···················· | ···················· | ···················· | ···················· | ···················· | ···················· | |
| H | 8538 | hCoV-19/England/LEED-2A8D98/2020\|EPI_ISL_539018\|20 | ···················· | ···················· | ···················· | ···················· | ···················· | ···················· | |
| H | 8539 | hCoV-19/England/LEED-2A8CB9/2020\|EPI_ISL_539020\|20 | ···················· | ···················· | ···················· | ···················· | ···················· | ···················· | |
| H | 8540 | hCoV-19/England/LEED-2A8AA0/2020\|EPI_ISL_539021\|20 | ···················· | ···················· | ···················· | ···················· | ···················· | ···················· | |
| H | 8541 | hCoV-19/England/LEED-2A9025/2020\|EPI_ISL_539022\|20 | ···················· | ···················· | ···················· | ···················· | ···················· | ···················· | |
| H | 8542 | hCoV-19/England/LEED-2A8949/2020\|EPI_ISL_539023\|20 | ···················· | ···················· | ···················· | ···················· | ···················· | ···················· | |
| H | 8543 | hCoV-19/England/LEED-2A908F/2020\|EPI_ISL_539024\|20 | ···················· | ···················· | ···················· | ···················· | ···················· | ···················· | |
| H | 8544 | hCoV-19/England/LEED-2A8985/2020\|EPI_ISL_539026\|20 | ···················· | ··················A· | ···················· | ···················· | ···················· | ···················· | 1 |
| H | 8545 | hCoV-19/England/LEED-2A8F38/2020\|EPI_ISL_539027\|20 | ···················· | ···A················ | ···················· | ···················· | ···················· | ···················· | |
| H | 8546 | hCoV-19/England/LEED-2A890D/2020\|EPI_ISL_539028\|20 | ···················· | ···················· | ···················· | ···················· | ···················· | ···················· | |
| H | 8547 | hCoV-19/England/LEED-2A81E7/2020\|EPI_ISL_539029\|20 | ···················· | ···················· | ···················· | ···················· | ···················· | ···················· | |
| H | 8548 | hCoV-19/England/LEED-2A85BE/2020\|EPI_ISL_539031\|20 | ···················· | ···················· | ···················· | ···················· | ···················· | ···················· | |
| H | 8549 | hCoV-19/England/LEED-2A884C/2020\|EPI_ISL_539032\|20 | ···················· | ···················· | ···················· | ···················· | ···················· | ···················· | |
| H | 8550 | hCoV-19/England/LEED-2A8D7A/2020\|EPI_ISL_539033\|20 | ···················· | ···················· | ···················· | ···················· | ···················· | ···················· | |
| H | 8551 | hCoV-19/England/LEED-2A8A0A/2020\|EPI_ISL_539034\|20 | ···················· | ···················· | ···················· | ···················· | ···················· | ···················· | |
| H | 8552 | hCoV-19/England/LEED-2A87E5/2020\|EPI_ISL_539036\|20 | ···················· | ···················· | ···················· | ···················· | ···················· | ···················· | |
| H | 8553 | hCoV-19/England/LEED-2A8BDA/2020\|EPI_ISL_539037\|20 | ···················· | ···················· | ···················· | ···················· | ···················· | ···················· | |
| H | 8554 | hCoV-19/England/LEED-2A8A55/2020\|EPI_ISL_539038\|20 | ···················· | ···················· | ···················· | ···················· | ···················· | ···················· | |
| H | 8555 | hCoV-19/England/LEED-2A8150/2020\|EPI_ISL_539039\|20 | ···················· | ···················· | ···················· | ···················· | ···················· | ···················· | |
| H | 8556 | hCoV-19/England/LEED-2A8A91/2020\|EPI_ISL_539040\|20 | ···················· | ···················· | ···················· | ···················· | ···················· | ···················· | |
| H | 8557 | hCoV-19/England/LEED-2A85FA/2020\|EPI_ISL_539041\|20 | ···················· | ···················· | ···················· | ···················· | ···················· | ···················· | |
| H | 8558 | hCoV-19/England/LEED-2A8F65/2020\|EPI_ISL_539043\|20 | ···················· | ···················· | ···················· | ···················· | ···················· | ···················· | |
| H | 8559 | hCoV-19/England/LEED-2A8ADD/2020\|EPI_ISL_539044\|20 | ···················· | ···················· | ···················· | ···················· | ···················· | ···················· | |
| H | 8560 | hCoV-19/England/LEED-2A8A73/2020\|EPI_ISL_539045\|20 | ···················· | ···················· | ···················· | ···················· | ···················· | ···················· | |
| H | 8561 | hCoV-19/England/LEED-2A8888/2020\|EPI_ISL_539047\|20 | ···················· | ···················· | ···················· | ···················· | ···················· | ···················· | |
| H | 8562 | hCoV-19/England/LEED-2A88A6/2020\|EPI_ISL_539051\|20 | ···················· | ···················· | ···················· | ···················· | ···················· | ···················· | |
| H | 8563 | hCoV-19/England/LEED-2A85EB/2020\|EPI_ISL_539053\|20 | ···················· | ···················· | ···················· | ···················· | ···················· | ···················· | |
| H | 8564 | hCoV-19/England/LEED-2A883D/2020\|EPI_ISL_539054\|20 | ···················· | ···················· | ···················· | ···················· | ···················· | ···················· | |
| H | 8565 | hCoV-19/England/LEED-2A83A5/2020\|EPI_ISL_539055\|20 | ···················· | ···················· | ···················· | ···················· | ···················· | ···················· | |
| H | 8566 | hCoV-19/England/LEED-2A8B8F/2020\|EPI_ISL_539058\|20 | ···················· | ···A················ | ···················· | ···················· | ···················· | ···················· | |
| H | 8567 | hCoV-19/England/QEUH-998BB8/2020\|EPI_ISL_540000\|20 | ···················· | ···················· | ···················· | ···················· | ···················· | ···················· | |
| H | 8568 | hCoV-19/England/QEUH-998796/2020\|EPI_ISL_540001\|20 | ···················· | ···················· | ···················· | ···················· | ···················· | ···················· | |
| H | 8569 | hCoV-19/England/QEUH-998839/2020\|EPI_ISL_540002\|20 | ···················· | ···················· | ···················· | ···················· | ···················· | ···················· | |
| H | 8570 | hCoV-19/England/QEUH-9984CC/2020\|EPI_ISL_540004\|20 | ···················· | ···················· | ···················· | ···················· | ·····T·············· | ···················· | |
| H | 8571 | hCoV-19/England/QEUH-98FDAF/2020\|EPI_ISL_540005\|20 | ···················· | ···················· | ···················· | ···················· | ···················· | ···················· | |
| H | 8572 | hCoV-19/England/QEUH-990084/2020\|EPI_ISL_540006\|20 | ···················· | ···················· | ···················· | ···················· | ···················· | ···················· | |
| H | 8573 | hCoV-19/England/QEUH-98FEF7/2020\|EPI_ISL_540007\|20 | ···················· | ···················· | ···················· | ···················· | ···················· | ···················· | |
| H | 8574 | hCoV-19/Scotland/QEUH-998383/2020\|EPI_ISL_540008\|2 | ···················· | ···················· | ···················· | ···················· | ···················· | ···················· | |
| H | 8575 | hCoV-19/England/QEUH-98F941/2020\|EPI_ISL_540009\|20 | ···················· | ···················· | ···················· | ···················· | ···················· | ···················· | |
| H | 8576 | hCoV-19/Scotland/QEUH-9901AF/2020\|EPI_ISL_540010\|2 | ···················· | ···················· | ···················· | ···················· | ···················· | ···················· | |
| H | 8577 | hCoV-19/England/QEUH-998CC4/2020\|EPI_ISL_540011\|20 | ···················· | ···················· | ···················· | ···················· | ···················· | ···················· | |
| H | 8578 | hCoV-19/England/QEUH-9983CF/2020\|EPI_ISL_540012\|20 | ···················· | ···················· | ···················· | ···················· | ···················· | ···················· | |
| H | 8579 | hCoV-19/England/QEUH-98F668/2020\|EPI_ISL_540013\|20 | ···················· | ···················· | ···················· | ···················· | ···················· | ···················· | |
| H | 8580 | hCoV-19/England/QEUH-9989AF/2020\|EPI_ISL_540014\|20 | ···················· | ···················· | ···················· | ···················· | ···················· | ···················· | |
| H | 8581 | hCoV-19/England/QEUH-9987D2/2020\|EPI_ISL_540016\|20 | ···················· | ·A·················· | ···················· | ···················· | ···················· | ···················· | |
| H | 8582 | hCoV-19/England/QEUH-998514/2020\|EPI_ISL_540017\|20 | ···················· | ···················· | ···················· | ···················· | ···················· | ···················· | |
| H | 8583 | hCoV-19/England/QEUH-998365/2020\|EPI_ISL_540018\|20 | ···················· | ···················· | ···················· | ···················· | ···················· | ···················· | |
| H | 8584 | hCoV-19/England/QEUH-98F56B/2020\|EPI_ISL_540019\|20 | ···················· | ···················· | ···················· | ···················· | ···T················ | ···················· | |
| H | 8585 | hCoV-19/England/QEUH-998AD9/2020\|EPI_ISL_540022\|20 | ···················· | ···················· | ···················· | ···················· | ···················· | ···················· | |

| | | | | | | | |
|---|---|---|---|---|---|---|---|
| H | 8586 hCoV-19/England/QEUH-98FB78/2020\|EPI_ISL_540023\|20 | .................... | .................... | ..................... | .................... | .................... | .................... |
| H | 8587 hCoV-19/England/QEUH-99038B/2020\|EPI_ISL_540025\|20 | .................... | .................... | ..................... | .................... | .................... | .................... |
| H | 8588 hCoV-19/England/QEUH-990057/2020\|EPI_ISL_540026\|20 | .................... | .................... | ..................... | .................... | .................... | .................... |
| H | 8589 hCoV-19/England/QEUH-99872D/2020\|EPI_ISL_540027\|20 | .................... | .................... | ..................... | .................... | .................... | .................... |
| H | 8590 hCoV-19/England/QEUH-990048/2020\|EPI_ISL_540028\|20 | .................... | .................... | ..................... | .................... | .................... | .................... |
| H | 8591 hCoV-19/England/QEUH-98F9F6/2020\|EPI_ISL_540029\|20 | .................... | .................... | ..................... | .................... | .....T.............. | .................... |
| H | 8592 hCoV-19/Wales/QEUH-998A42/2020\|EPI_ISL_540031\|2020 | .................... | .................... | ..................... | .................... | .................... | .................... |
| H | 8593 hCoV-19/England/QEUH-98F659/2020\|EPI_ISL_540032\|20 | .................... | .................... | ..................... | .................... | .................... | .................... |
| H | 8594 hCoV-19/England/QEUH-998453/2020\|EPI_ISL_540033\|20 | .................... | .................... | ..................... | .................... | .................... | .................... |
| H | 8595 hCoV-19/England/QEUH-998A24/2020\|EPI_ISL_540034\|20 | .................... | .................... | ..................... | .................... | .................... | .................... |
| H | 8596 hCoV-19/Scotland/QEUH-998523/2020\|EPI_ISL_540035\|2 | .................... | .................... | ..................... | .................... | .................... | .................... |
| H | 8597 hCoV-19/England/QEUH-9987F0/2020\|EPI_ISL_540036\|20 | .................... | .................... | ..................... | .................... | .................... | .................... |
| H | 8598 hCoV-19/England/QEUH-998B12/2020\|EPI_ISL_540037\|20 | .................... | .................... | ..................... | .................... | .................... | .................... |
| H | 8599 hCoV-19/England/QEUH-998954/2020\|EPI_ISL_540038\|20 | .................... | .................... | ..................... | .................... | .................... | .................... |
| H | 8600 hCoV-19/Scotland/QEUH-990190/2020\|EPI_ISL_540039\|2 | .................... | .................... | ..................... | .................... | .................... | .................... |
| H | 8601 hCoV-19/Scotland/QEUH-998CE2/2020\|EPI_ISL_540040\|2 | .................... | .................... | ..................... | .................... | .................... | .................... |
| H | 8602 hCoV-19/England/QEUH-998505/2020\|EPI_ISL_540041\|20 | .................... | .................... | ..................... | .................... | .................... | .................... |
| H | 8603 hCoV-19/England/QEUH-998480/2020\|EPI_ISL_540042\|20 | .................... | .................... | ..................... | .................... | .................... | .................... |
| H | 8604 hCoV-19/England/QEUH-98FB69/2020\|EPI_ISL_540043\|20 | .................... | .................... | ..................... | .................... | .................... | .................... |
| H | 8605 hCoV-19/Scotland/QEUH-98F6FF/2020\|EPI_ISL_540044\|2 | .................... | .................... | ..................... | .................... | .................... | .................... |
| H | 8606 hCoV-19/England/QEUH-998981/2020\|EPI_ISL_540045\|20 | .................... | .................... | ..................... | .................... | .................... | .................... |
| H | 8607 hCoV-19/England/QEUH-98FAF3/2020\|EPI_ISL_540046\|20 | .................... | .................... | ..................... | .................... | .................... | .................... |
| H | 8608 hCoV-19/USA/FL-BPHL-1378/2020\|EPI_ISL_541315\|2020- | .................... | .................... | ..................... | .................... | .................... | .................... |
| H | 8609 hCoV-19/USA/FL-BPHL-1381/2020\|EPI_ISL_541318\|2020- | .................... | .................... | ..................... | .................... | .................... | .................... |
| H | 8610 hCoV-19/USA/FL-BPHL-1382/2020\|EPI_ISL_541319\|2020- | .................... | .................... | ..................... | .................... | .................... | .................... |
| H | 8611 hCoV-19/USA/FL-BPHL-1386/2020\|EPI_ISL_541323\|2020- | .................... | .................... | ..................... | .................... | .................... | .................... |
| H | 8612 hCoV-19/USA/FL-BPHL-1394/2020\|EPI_ISL_541331\|2020- | .................... | .................... | ..................... | .................... | .................... | .................... |
| H | 8613 hCoV-19/Czech Republic/NRL-6833/2020\|EPI_ISL_54133 | .................... | .................... | ..................... | .................... | .................... | .................... |
| H | 8614 hCoV-19/Czech Republic/NRL-6834/2020\|EPI_ISL_54133 | .................... | .................... | ..................... | .................... | .................... | .................... |
| H | 8615 hCoV-19/Czech Republic/NRL-6835/2020\|EPI_ISL_54133 | .................... | .................... | ..................... | .................... | .................... | .................... |
| H | 8616 hCoV-19/Czech Republic/NRL-6849-8/2020\|EPI_ISL_541 | .................... | .................... | ..................... | .................... | .................... | .................... |
| H | 8617 hCoV-19/Czech Republic/NRL-7913/2020\|EPI_ISL_54133 | .................... | .................... | ..................... | .................... | .................... | .................... |
| H | 8618 hCoV-19/Czech Republic/NRL-7915/2020\|EPI_ISL_54133 | .................... | .................... | ..................... | .................... | .................... | .................... |
| H | 8619 hCoV-19/Brazil/PR-5617/2020\|EPI_ISL_541340\|2020-03 | .................... | .................... | ..................... | .................... | .................... | .................... |
| H | 8620 hCoV-19/Brazil/PR-5618/2020\|EPI_ISL_541341\|2020-03 | .................... | .................... | ..................... | .................... | .................... | .................... |
| H | 8621 hCoV-19/Brazil/PR-5619/2020\|EPI_ISL_541342\|2020-03 | .................... | .................... | ..................... | .................... | .................... | .................... |
| H | 8622 hCoV-19/Brazil/PR-5620/2020\|EPI_ISL_541343\|2020-03 | .................... | .................... | ..................... | .................... | .................... | .................... |
| H | 8623 hCoV-19/Brazil/PR-5621/2020\|EPI_ISL_541344\|2020-03 | .................... | .................... | ..................... | .................... | .................... | .................... |
| H | 8624 hCoV-19/Brazil/PR-5622/2020\|EPI_ISL_541345\|2020-03 | .................... | .................... | ..................... | .................... | .................... | .................... |
| H | 8625 hCoV-19/Brazil/PR-5623/2020\|EPI_ISL_541346\|2020-03 | .................... | .................... | ..................... | .................... | .................... | .................... |
| H | 8626 hCoV-19/Brazil/RJ-1783/2020\|EPI_ISL_541347\|2020-04 | .................... | .................... | ..................... | .................... | .................... | .................... |
| H | 8627 hCoV-19/Brazil/RJ-1798/2020\|EPI_ISL_541348\|2020-04 | .................... | .................... | ..................... | .................... | .................... | .................... |
| H | 8628 hCoV-19/Brazil/RJ-1830/2020\|EPI_ISL_541349\|2020-04 | .................... | .................... | ..................... | .................... | .................... | .................... |
| H | 8629 hCoV-19/Brazil/RJ-1862/2020\|EPI_ISL_541350\|2020-04 | .................... | .................... | ..................... | .................... | .................... | .................... |
| H | 8630 hCoV-19/Brazil/RJ-1870/2020\|EPI_ISL_541351\|2020-04 | .................... | .................... | ..................... | .................... | .................... | .................... |
| H | 8631 hCoV-19/Brazil/RJ-1953/2020\|EPI_ISL_541352\|2020-04 | .................... | .................... | ..................... | .................... | .................... | .................... |
| H | 8632 hCoV-19/Brazil/RJ-2100/2020\|EPI_ISL_541353\|2020-04 | .................... | .................... | ..................... | .................... | .................... | .................... |
| H | 8633 hCoV-19/Brazil/RJ-2155/2020\|EPI_ISL_541354\|2020-04 | .................... | .................... | ..................... | .................... | .................... | .................... |
| H | 8634 hCoV-19/Brazil/RJ-2192/2020\|EPI_ISL_541355\|2020-04 | .................... | .................... | ..................... | .................... | .................... | .................... |
| H | 8635 hCoV-19/Brazil/RJ-2231/2020\|EPI_ISL_541356\|2020-04 | .................... | .................... | ..................... | .................... | .................... | .................... |
| H | 8636 hCoV-19/Brazil/RJ-2342/2020\|EPI_ISL_541357\|2020-04 | .................... | .................... | ..................... | .................... | .................... | .................... |
| H | 8637 hCoV-19/Brazil/RJ-2343/2020\|EPI_ISL_541358\|2020-04 | .................... | .................... | ..................... | .................... | .................... | .................... |
| H | 8638 hCoV-19/Brazil/RJ-2369/2020\|EPI_ISL_541359\|2020-04 | .................... | .................... | ..................... | .................... | .................... | .................... |
| H | 8639 hCoV-19/Brazil/RJ-2376/2020\|EPI_ISL_541360\|2020-04 | .................... | .................... | ..................... | .................... | .................... | .................... |
| H | 8640 hCoV-19/Brazil/RJ-2377/2020\|EPI_ISL_541361\|2020-04 | .................... | .................... | ..................... | .................... | .................... | .................... |
| H | 8641 hCoV-19/Brazil/RJ-2903/2020\|EPI_ISL_541362\|2020-04 | .................... | .................... | ..................... | .................... | .................... | .................... |
| H | 8642 hCoV-19/Brazil/RJ-2904/2020\|EPI_ISL_541363\|2020-04 | .................... | .................... | ..................... | .................... | .................... | .................... |
| H | 8643 hCoV-19/Brazil/RJ-2938/2020\|EPI_ISL_541364\|2020-04 | .................... | .................... | ..................... | .................... | .................... | .................... |
| H | 8644 hCoV-19/Brazil/RJ-2950/2020\|EPI_ISL_541365\|2020-04 | .................... | .................... | ..................... | .................... | .................... | .................... |
| H | 8645 hCoV-19/Brazil/RJ-2951/2020\|EPI_ISL_541366\|2020-04 | .................... | .................... | ..................... | .................... | .................... | .................... |
| H | 8646 hCoV-19/Brazil/RJ-2966/2020\|EPI_ISL_541367\|2020-04 | .................... | .................... | ..................... | .................... | .................... | .................... |
| H | 8647 hCoV-19/Brazil/RJ-2999/2020\|EPI_ISL_541368\|2020-04 | .................... | .................... | ..................... | .................... | .................... | .................... |
| H | 8648 hCoV-19/Brazil/RJ-3021/2020\|EPI_ISL_541369\|2020-04 | .................... | .................... | ..................... | .................... | .................... | .................... |
| H | 8649 hCoV-19/Brazil/SC-770/2020\|EPI_ISL_541370\|2020-03- | .................... | .................... | ..................... | .................... | .................... | .................... |
| H | 8650 hCoV-19/Brazil/SE-6529/2020\|EPI_ISL_541372\|2020-03 | .................... | .................... | ..................... | .................... | .................... | .................... |
| H | 8651 hCoV-19/Brazil/SE-6530/2020\|EPI_ISL_541373\|2020-03 | .................... | .................... | ..................... | .................... | .................... | .................... |
| H | 8652 hCoV-19/Brazil/SE-6533/2020\|EPI_ISL_541374\|2020-03 | .................... | .................... | ..................... | .................... | .................... | .................... |
| H | 8653 hCoV-19/Brazil/SE-6535/2020\|EPI_ISL_541375\|2020-03 | .................... | .................... | ..................... | .................... | .................... | .................... |
| H | 8654 hCoV-19/Brazil/SE-6536/2020\|EPI_ISL_541376\|2020-03 | .................... | .................... | ..................... | .................... | .................... | .................... |
| H | 8655 hCoV-19/Brazil/SE-6539/2020\|EPI_ISL_541377\|2020-04 | .................... | .................... | ..................... | .................... | .................... | .................... |
| H | 8656 hCoV-19/USA/TX-HMH-1309/2020\|EPI_ISL_542643\|2020-0 | .................... | .................... | ..................... | .................... | .................... | .................... |
| H | 8657 hCoV-19/USA/TX-HMH-1310/2020\|EPI_ISL_542644\|2020-0 | .................... | .................... | ..................... | .................... | .................... | .................... |
| H | 8658 hCoV-19/USA/TX-HMH-1311/2020\|EPI_ISL_542645\|2020-0 | .................... | .................... | ..................... | .................... | .................... | .................... |
| H | 8659 hCoV-19/USA/TX-HMH-1313/2020\|EPI_ISL_542646\|2020-0 | .................... | .................... | ..................... | .................... | .................... | .................... |
| H | 8660 hCoV-19/USA/TX-HMH-1315/2020\|EPI_ISL_542647\|2020-0 | .................... | .................... | ..................... | .................... | .................... | .................... |
| H | 8661 hCoV-19/USA/TX-HMH-1316/2020\|EPI_ISL_542648\|2020-0 | .................... | .................... | ..................... | .................... | .................... | .................... |
| H | 8662 hCoV-19/USA/TX-HMH-1317/2020\|EPI_ISL_542649\|2020-0 | .................... | .................... | ..................... | .................... | .................... | .................... |
| H | 8663 hCoV-19/USA/TX-HMH-1319/2020\|EPI_ISL_542651\|2020-0 | .................... | .................... | ..................... | .................... | .................... | .................... |
| H | 8664 hCoV-19/USA/TX-HMH-1322/2020\|EPI_ISL_542652\|2020-0 | .................... | .................... | ..................... | .................... | .................... | .................... |
| H | 8665 hCoV-19/USA/TX-HMH-1325/2020\|EPI_ISL_542653\|2020-0 | .................... | .................... | ..................... | .................... | .................... | .................... |

| | | | | | | | |
|---|---|---|---|---|---|---|---|
| H | 8666 hCoV-19/USA/TX-HMH-1332/2020\|EPI_ISL_542654\|2020-0 | ···················· | ···················· | ···················· | ···················· | ···················· | N··················· |
| H | 8667 hCoV-19/USA/TX-HMH-1334/2020\|EPI_ISL_542655\|2020-0 | ···················· | ···················· | ···················· | ···················· | ···················· | ···················· |
| H | 8668 hCoV-19/USA/TX-HMH-1337/2020\|EPI_ISL_542657\|2020-0 | ···················· | ···················· | ···················· | ···················· | ···················· | ···················· |
| H | 8669 hCoV-19/USA/TX-HMH-1338/2020\|EPI_ISL_542658\|2020-0 | ···················· | ···················· | ···················· | ···················· | ···················· | ···················· |
| H | 8671 hCoV-19/USA/TX-HMH-1342/2020\|EPI_ISL_542660\|2020-0 | ···················· | ···················· | ···················· | ···················· | ···················· | ···················· |
| H | 8672 hCoV-19/USA/TX-HMH-1348/2020\|EPI_ISL_542663\|2020-0 | ···················· | ···················· | ···················· | ···················· | ···················· | ···················· |
| H | 8674 hCoV-19/USA/TX-HMH-1351/2020\|EPI_ISL_542665\|2020-0 | ···················· | ···················· | ···················· | ···················· | ···················· | ···················· |
| H | 8675 hCoV-19/USA/TX-HMH-1353/2020\|EPI_ISL_542666\|2020-0 | ···················· | ···················· | ···················· | ···················· | ···················· | ···················· |
| H | 8676 hCoV-19/USA/TX-HMH-1354/2020\|EPI_ISL_542667\|2020-0 | ···················· | ···················· | ···················· | ···················· | ···················· | ···················· |
| H | 8677 hCoV-19/USA/TX-HMH-1358/2020\|EPI_ISL_542668\|2020-0 | ···················· | ···················· | ···················· | ···················· | ···················· | ···················· |
| H | 8678 hCoV-19/USA/TX-HMH-1359/2020\|EPI_ISL_542669\|2020-0 | ···················· | ···················· | ···················· | ···················· | ···················· | ···················· |
| H | 8679 hCoV-19/USA/TX-HMH-1362/2020\|EPI_ISL_542671\|2020-0 | ···················· | ···················· | ···················· | ···················· | ···················· | ···················· |
| H | 8680 hCoV-19/USA/TX-HMH-1364/2020\|EPI_ISL_542673\|2020-0 | ···················· | ···················· | ···················· | ···················· | ···················· | ···················· |
| H | 8681 hCoV-19/USA/TX-HMH-1365/2020\|EPI_ISL_542674\|2020-0 | ···················· | ···················· | ···················· | ···················· | ···················· | ···················· |
| H | 8682 hCoV-19/USA/TX-HMH-1366/2020\|EPI_ISL_542675\|2020-0 | ···················· | ···················· | ···················· | ···················· | ···················· | ···················· |
| H | 8683 hCoV-19/USA/TX-HMH-1369/2020\|EPI_ISL_542676\|2020-0 | ···················· | ···················· | ···················· | ···················· | ···················· | ···················· |
| H | 8684 hCoV-19/USA/TX-HMH-1370/2020\|EPI_ISL_542677\|2020-0 | ···················· | ···················· | ···················· | ···················· | ···················· | ···················· |
| H | 8685 hCoV-19/USA/TX-HMH-1371/2020\|EPI_ISL_542678\|2020-0 | ···················· | ···················· | ···················· | ···················· | ···················· | ···················· |
| H | 8687 hCoV-19/USA/TX-HMH-1374/2020\|EPI_ISL_542680\|2020-0 | ···················· | ···················· | ···················· | ···················· | ···T················ | ···················· |
| H | 8688 hCoV-19/USA/TX-HMH-1375/2020\|EPI_ISL_542681\|2020-0 | ···················· | ···················· | ···················· | ···················· | ···················· | ···················· |
| H | 8689 hCoV-19/USA/TX-HMH-1376/2020\|EPI_ISL_542682\|2020-0 | ···················· | ···················· | ···················· | ···················· | ···················· | ···················· |
| H | 8690 hCoV-19/USA/TX-HMH-1377/2020\|EPI_ISL_542683\|2020-0 | ···················· | ···················· | ···················· | ···················· | ···················· | ···················· |
| H | 8691 hCoV-19/USA/TX-HMH-1380/2020\|EPI_ISL_542684\|2020-0 | ···················· | ···················· | ···················· | ···················· | ···················· | ···················· |
| H | 8692 hCoV-19/USA/TX-HMH-1381/2020\|EPI_ISL_542685\|2020-0 | ···················· | ···················· | ···················· | ···················· | ···················· | ···················· |
| H | 8693 hCoV-19/USA/TX-HMH-1384/2020\|EPI_ISL_542687\|2020-0 | ···················· | ···················· | ···················· | ···················· | ···················· | ···················· |
| H | 8694 hCoV-19/USA/TX-HMH-1385/2020\|EPI_ISL_542688\|2020-0 | ···················· | ···················· | ···················· | ···················· | ···················· | ···················· |
| H | 8695 hCoV-19/USA/TX-HMH-1386/2020\|EPI_ISL_542689\|2020-0 | ···················· | ···················· | ···················· | ···················· | ···················· | ···················· |
| H | 8696 hCoV-19/USA/TX-HMH-1388/2020\|EPI_ISL_542690\|2020-0 | ···················· | ···················· | ···················· | ···················· | ···················· | ···················· |
| H | 8697 hCoV-19/USA/TX-HMH-1389/2020\|EPI_ISL_542691\|2020-0 | ···················· | ···················· | ···················· | ···················· | ···················· | ···················· |
| H | 8698 hCoV-19/USA/TX-HMH-1391/2020\|EPI_ISL_542692\|2020-0 | ···················· | ···················· | ···················· | ···················· | ···················· | ···················· |
| H | 8699 hCoV-19/USA/TX-HMH-1392/2020\|EPI_ISL_542693\|2020-0 | ···················· | ···················· | ···················· | ···················· | ···················· | ···················· |
| H | 8700 hCoV-19/USA/TX-HMH-1396/2020\|EPI_ISL_542694\|2020-0 | ···················· | ···················· | ···················· | ···················· | ···················· | ···················· |
| H | 8701 hCoV-19/USA/TX-HMH-1397/2020\|EPI_ISL_542695\|2020-0 | ···················· | ···················· | ···················· | ···················· | ···················· | ···················· |
| H | 8702 hCoV-19/USA/TX-HMH-1398/2020\|EPI_ISL_542696\|2020-0 | ···················· | ···················· | ···················· | ···················· | ···················· | ···················· |
| H | 8703 hCoV-19/USA/TX-HMH-1400/2020\|EPI_ISL_542698\|2020-0 | ···················· | ···················· | ···················· | ···················· | ···················· | ···················· |
| H | 8704 hCoV-19/USA/TX-HMH-1401/2020\|EPI_ISL_542699\|2020-0 | ···················· | ···················· | ···················· | ···················· | ···················· | ···················· |
| H | 8705 hCoV-19/Scotland/QEUH-96E16E/2020\|EPI_ISL_530488\|2 | ···················· | ···················· | ···················· | ···················· | ···················· | ···················· |
| H | 8706 hCoV-19/Scotland/QEUH-96E83C/2020\|EPI_ISL_530489\|2 | ···················· | ···················· | ···················· | ···················· | ···················· | ···················· |
| H | 8707 hCoV-19/England/QEUH-96F570/2020\|EPI_ISL_530490\|20 | ···················· | ···················· | ···················· | ···················· | ···················· | ···················· |
| H | 8708 hCoV-19/England/QEUH-96F464/2020\|EPI_ISL_530491\|20 | ···················· | ···················· | ···················· | ···················· | ···················· | ···················· |
| H | 8709 hCoV-19/England/QEUH-96F19A/2020\|EPI_ISL_530493\|20 | ···················· | ·A·················· | ···················· | ···················· | ···················· | ···················· |
| H | 8710 hCoV-19/England/QEUH-96F24C/2020\|EPI_ISL_530494\|20 | ···················· | ···················· | ···················· | ···················· | ···················· | ···················· |
| H | 8711 hCoV-19/England/QEUH-96F3B2/2020\|EPI_ISL_530495\|20 | ···················· | ···················· | ···················· | ···················· | ···················· | ···················· |
| H | 8712 hCoV-19/England/QEUH-96E3D1/2020\|EPI_ISL_530496\|20 | ···················· | ···················· | ···················· | ···················· | ···················· | ···················· |
| H | 8713 hCoV-19/England/QEUH-96EE3A/2020\|EPI_ISL_530498\|20 | ···················· | ···················· | ···················· | ···················· | ···················· | ···················· |
| H | 8714 hCoV-19/England/QEUH-96F507/2020\|EPI_ISL_530499\|20 | ···················· | ···················· | ···················· | ···················· | ···················· | ···················· |
| H | 8715 hCoV-19/Northern Ireland/QEUH-96E5F9/2020\|EPI_ISL_ | ···················· | ···················· | ···················· | ···················· | ···················· | ···················· |
| H | 8716 hCoV-19/Northern Ireland/QEUH-96EA72/2020\|EPI_ISL_ | ···················· | ···················· | ···················· | ···················· | ···················· | ···················· |
| H | 8717 hCoV-19/England/QEUH-96F67D/2020\|EPI_ISL_530612\|20 | ···················· | ···················· | ···················· | ···················· | ···················· | ···················· |
| H | 8718 hCoV-19/England/QEUH-96E061/2020\|EPI_ISL_530613\|20 | ···················· | ···················· | ···················· | ···················· | ···················· | ···················· |
| H | 8719 hCoV-19/England/QEUH-96E82D/2020\|EPI_ISL_530615\|20 | ···················· | ·A·················· | ···················· | ···················· | ···················· | ···················· |
| H | 8720 hCoV-19/England/QEUH-96F1F4/2020\|EPI_ISL_530616\|20 | ···················· | ···················· | ···················· | ···················· | ···················· | ···················· |
| H | 8721 hCoV-19/Northern Ireland/QEUH-96EAAF/2020\|EPI_ISL_ | ···················· | ···················· | ···················· | ···················· | ···················· | ···················· |
| H | 8722 hCoV-19/England/QEUH-96E09E/2020\|EPI_ISL_530618\|20 | ···················· | ···················· | ···················· | ···················· | ···················· | ···················· |
| H | 8723 hCoV-19/Northern Ireland/QEUH-96F0CA/2020\|EPI_ISL_ | ···················· | ···················· | ···················· | ···················· | ···················· | ···················· |
| H | 8724 hCoV-19/England/QEUH-96EA09/2020\|EPI_ISL_530620\|20 | ···················· | ···················· | ···················· | ···················· | ···················· | ···················· |
| H | 8725 hCoV-19/Northern Ireland/QEUH-96EE1C/2020\|EPI_ISL_ | ···················· | ···················· | ···················· | ···················· | ···················· | ···················· |
| H | 8726 hCoV-19/Northern Ireland/QEUH-96D044/2020\|EPI_ISL_ | ···················· | ···················· | ···················· | ···················· | ···················· | ···················· |
| H | 8727 hCoV-19/Northern Ireland/QEUH-96D624/2020\|EPI_ISL_ | ···················· | ···················· | ···················· | ···················· | ···················· | ···················· |
| H | 8728 hCoV-19/Northern Ireland/QEUH-96D633/2020\|EPI_ISL_ | ···················· | ···················· | ···················· | ···················· | ···················· | ···················· |
| H | 8729 hCoV-19/Northern Ireland/QEUH-96D8B5/2020\|EPI_ISL_ | ···················· | ···················· | ···················· | ···················· | ···················· | ···················· |
| H | 8730 hCoV-19/England/QEUH-96DF38/2020\|EPI_ISL_530628\|20 | ···················· | ···················· | ···················· | ···················· | ···················· | ···················· |
| H | 8731 hCoV-19/England/QEUH-96D19C/2020\|EPI_ISL_530629\|20 | ···················· | ···················· | ···················· | ···················· | ···················· | ···················· |
| H | 8732 hCoV-19/England/QEUH-96D1BA/2020\|EPI_ISL_530631\|20 | ···················· | ···················· | ···················· | ···················· | ···················· | ···················· |
| H | 8733 hCoV-19/Northern Ireland/QEUH-96CEA5/2020\|EPI_ISL_ | ···················· | ···················· | ···················· | ···················· | ···················· | ···················· |
| H | 8734 hCoV-19/Northern Ireland/QEUH-96CC8D/2020\|EPI_ISL_ | ···················· | ···················· | ···················· | ···················· | ···················· | ···················· |
| H | 8735 hCoV-19/Northern Ireland/QEUH-96DD2F/2020\|EPI_ISL_ | ···················· | ···················· | ···················· | ···················· | ···················· | ···················· |
| H | 8736 hCoV-19/England/QEUH-96D2D5/2020\|EPI_ISL_530635\|20 | ···················· | ·A·················· | ···················· | ···················· | ···················· | ···················· |
| H | 8737 hCoV-19/Northern Ireland/QEUH-96CCC9/2020\|EPI_ISL_ | ···················· | ···················· | ···················· | ···················· | ···················· | ···················· |
| H | 8738 hCoV-19/Northern Ireland/QEUH-96DACE/2020\|EPI_ISL_ | ···················· | ···················· | ···················· | ···················· | ···················· | ···················· |
| H | 8739 hCoV-19/Northern Ireland/QEUH-96D053/2020\|EPI_ISL_ | ···················· | ···················· | ···················· | ···················· | ···················· | ···················· |
| H | 8740 hCoV-19/England/QEUH-96CB08/2020\|EPI_ISL_530641\|20 | ···················· | ···················· | ···················· | ···················· | ···················· | ···················· |
| H | 8741 hCoV-19/England/QEUH-9603EE/2020\|EPI_ISL_531942\|20 | ···················· | ···················· | ···················· | ···················· | ···················· | ···················· |
| H | 8742 hCoV-19/Scotland/QEUH-960F80/2020\|EPI_ISL_531943\|2 | ···················· | ···················· | ···················· | ···················· | ···················· | ···················· |
| H | 8743 hCoV-19/England/QEUH-96082B/2020\|EPI_ISL_531944\|20 | ···················· | ···················· | ···················· | ···················· | ···················· | ···················· |
| H | 8744 hCoV-19/Scotland/QEUH-9610B9/2020\|EPI_ISL_531945\|2 | ···················· | ···················· | ···················· | ···················· | ···················· | ···················· |
| H | 8745 hCoV-19/Northern Ireland/QEUH-960DE0/2020\|EPI_ISL_ | ···················· | ···················· | ···················· | ···················· | ···················· | ···················· |
| H | 8746 hCoV-19/England/QEUH-960A16/2020\|EPI_ISL_531948\|20 | ···················· | ···················· | ···················· | ···················· | ···················· | ···················· |
| H | 8747 hCoV-19/England/QEUH-9602D2/2020\|EPI_ISL_531949\|20 | ···················· | ···················· | ···················· | ···················· | ···················· | ···················· |
| H | 8748 hCoV-19/England/QEUH-9605E8/2020\|EPI_ISL_531950\|20 | ···················· | ···················· | ···················· | ···················· | ···················· | ···················· |

| | | | | | | | | |
|---|---|---|---|---|---|---|---|---|
| H | 8749 | hCoV-19/Scotland/QEUH-9610AA/2020\|EPI_ISL_531951\|2 | ···················· | ···················· | ····················· | ···················· | ···················· | ···················· |
| H | 8750 | hCoV-19/England/QEUH-96080D/2020\|EPI_ISL_531952\|20 | ···················· | ···················· | ····················· | ···················· | ···················· | ···················· |
| H | 8751 | hCoV-19/England/QEUH-96071F/2020\|EPI_ISL_531953\|20 | ···················· | ···················· | ····················· | ···················· | ···················· | ···················· |
| H | 8752 | hCoV-19/Scotland/QEUH-9610C8/2020\|EPI_ISL_531954\|2 | ···················· | ···················· | ····················· | ···················· | ···················· | ···················· |
| H | 8753 | hCoV-19/England/QEUH-960F71/2020\|EPI_ISL_531955\|20 | ···················· | ···················· | ····················· | ···················· | ···················· | ···················· |
| H | 8754 | hCoV-19/England/QEUH-9609FB/2020\|EPI_ISL_531956\|20 | ···················· | ·A·······A·········· | ····················· | ···················· | ···················· | ···················· |
| H | 8755 | hCoV-19/England/QEUH-96059D/2020\|EPI_ISL_531957\|20 | ···················· | ···················· | ····················· | ···················· | ···················· | ···················· |
| H | 8756 | hCoV-19/Northern Ireland/QEUH-9607E2/2020\|EPI_ISL_ | ···················· | ···················· | ····················· | ···················· | ···················· | ···················· |
| H | 8757 | hCoV-19/England/QEUH-960339/2020\|EPI_ISL_531959\|20 | ···················· | ···················· | ····················· | ···················· | ···················· | ···················· |
| H | 8758 | hCoV-19/England/QEUH-96107D/2020\|EPI_ISL_531960\|20 | ···················· | ···················· | ····················· | ···················· | ···················· | ···················· |
| H | 8759 | hCoV-19/England/QEUH-9605CA/2020\|EPI_ISL_531961\|20 | ···················· | ···················· | ····················· | ···················· | ···················· | ···················· |
| H | 8760 | hCoV-19/England/QEUH-960B40/2020\|EPI_ISL_531962\|20 | ···················· | ···················· | ····················· | ···················· | ···················· | ···················· |
| H | 8761 | hCoV-19/Scotland/QEUH-96131A/2020\|EPI_ISL_531963\|2 | ···················· | ···················· | ····················· | ···················· | ···················· | ···················· |
| H | 8762 | hCoV-19/England/QEUH-96013F/2020\|EPI_ISL_531964\|20 | ···················· | ···················· | ···········T········· | ···················· | ···················· | ···················· |
| H | 8763 | hCoV-19/England/QEUH-9603FD/2020\|EPI_ISL_531965\|20 | ···················· | ···················· | ····················· | ···················· | ···················· | ···················· |
| H | 8764 | hCoV-19/England/QEUH-960FCC/2020\|EPI_ISL_531966\|20 | ···················· | ···················· | ····················· | ···················· | ···················· | ···················· |
| H | 8765 | hCoV-19/England/QEUH-9612A4/2020\|EPI_ISL_531967\|20 | ···················· | ···················· | ····················· | ···················· | ···················· | ···················· |
| H | 8766 | hCoV-19/England/QEUH-960621/2020\|EPI_ISL_531968\|20 | ···················· | ···················· | ····················· | ···················· | ···················· | ···················· |
| H | 8767 | hCoV-19/England/QEUH-960A07/2020\|EPI_ISL_531969\|20 | ···················· | ···················· | ····················· | ···················· | ···················· | ···················· |
| H | 8768 | hCoV-19/England/QEUH-960FDB/2020\|EPI_ISL_531970\|20 | ···················· | ···················· | ····················· | ···················· | ···················· | ···················· |
| H | 8769 | hCoV-19/England/QEUH-960EDE/2020\|EPI_ISL_531971\|20 | ···················· | ···················· | ····················· | ···················· | ···················· | ···················· |
| H | 8770 | hCoV-19/England/QEUH-960ACB/2020\|EPI_ISL_531972\|20 | ···················· | ···················· | ····················· | ···················· | ···················· | ···················· |
| H | 8771 | hCoV-19/USA/NM-NMDOH-2020230022/2020\|EPI_ISL_53530 | ···················· | ····T··············· | ····················· | ···················· | ···················· | ···················· |
| H | 8772 | hCoV-19/USA/NM-NMDOH-2020230030/2020\|EPI_ISL_53530 | ···················· | ···················· | ····················· | ···················· | ···················· | ···················· |
| H | 8773 | hCoV-19/USA/NM-NMDOH-2020230061/2020\|EPI_ISL_53530 | ···················· | ···················· | ····················· | ···················· | ···················· | ···················· |
| H | 8774 | hCoV-19/USA/NM-NMDOH-2020230064/2020\|EPI_ISL_53530 | ···················· | ···················· | ····················· | ···················· | ···················· | ···················· |
| H | 8775 | hCoV-19/USA/LA-CDC-1264/2020\|EPI_ISL_535305\|2020-0 | ···················· | ···················· | ····················· | ···················· | ···················· | ···················· |
| H | 8776 | hCoV-19/USA/LA-CDC-1265/2020\|EPI_ISL_535306\|2020-0 | ···················· | ···················· | ····················· | ···················· | ···················· | ···················· |
| H | 8777 | hCoV-19/USA/LA-CDC-1266/2020\|EPI_ISL_535307\|2020-0 | ···················· | ···················· | ····················· | ···················· | ···················· | ···················· |
| H | 8778 | hCoV-19/USA/LA-CDC-1267/2020\|EPI_ISL_535308\|2020-0 | ···················· | ···················· | ····················· | ···················· | ···················· | ···················· |
| H | 8779 | hCoV-19/USA/LA-CDC-1268/2020\|EPI_ISL_535309\|2020-0 | ···················· | ···················· | ····················· | ···················· | ···················· | ···················· |
| H | 8780 | hCoV-19/USA/LA-CDC-1269/2020\|EPI_ISL_535310\|2020-0 | ···················· | ···················· | ····················· | ···················· | ···················· | ···················· |
| H | 8781 | hCoV-19/USA/LA-CDC-1270/2020\|EPI_ISL_535311\|2020-0 | ···················· | ···················· | ····················· | ···················· | ···················· | ···················· |
| H | 8782 | hCoV-19/USA/LA-CDC-1271/2020\|EPI_ISL_535312\|2020-0 | ···················· | ···················· | ····················· | ···················· | ···················· | ···················· |
| H | 8783 | hCoV-19/USA/LA-CDC-1272/2020\|EPI_ISL_535313\|2020-0 | ···················· | ···················· | ····················· | ···················· | ···················· | ···················· |
| H | 8784 | hCoV-19/USA/LA-CDC-1273/2020\|EPI_ISL_535314\|2020-0 | ···················· | ···················· | ····················· | ···················· | ···················· | ···················· |
| H | 8785 | hCoV-19/USA/LA-CDC-1274/2020\|EPI_ISL_535315\|2020-0 | ···················· | ···················· | ····················· | ···················· | ···················· | ···················· |
| H | 8786 | hCoV-19/USA/LA-CDC-1275/2020\|EPI_ISL_535316\|2020-0 | ···················· | ···················· | ····················· | ···················· | ···················· | ···················· |
| H | 8787 | hCoV-19/USA/LA-CDC-1276/2020\|EPI_ISL_535317\|2020-0 | ···················· | ···················· | ····················· | ···················· | ···················· | ···················· |
| H | 8788 | hCoV-19/USA/LA-CDC-1277/2020\|EPI_ISL_535318\|2020-0 | ···················· | ···················· | ····················· | ···················· | ···················· | ···················· |
| H | 8789 | hCoV-19/USA/LA-CDC-1278/2020\|EPI_ISL_535319\|2020-0 | ···················· | ···················· | ····················· | ···················· | ···················· | ···················· |
| H | 8790 | hCoV-19/USA/LA-CDC-1279/2020\|EPI_ISL_535320\|2020-0 | ···················· | ···················· | ····················· | ···················· | ···················· | ···················· |
| H | 8791 | hCoV-19/USA/LA-CDC-1280/2020\|EPI_ISL_535321\|2020-0 | ···················· | ···················· | ····················· | ···················· | ···················· | ···················· |
| H | 8792 | hCoV-19/USA/LA-CDC-1281/2020\|EPI_ISL_535322\|2020-0 | ···················· | ···················· | ····················· | ···················· | ···················· | ···················· |
| H | 8793 | hCoV-19/USA/LA-CDC-1282/2020\|EPI_ISL_535323\|2020-0 | ···················· | ···················· | ····················· | ···················· | ···················· | ···················· |
| H | 8794 | hCoV-19/USA/LA-CDC-1283/2020\|EPI_ISL_535324\|2020-0 | ···················· | ···················· | ····················· | ···················· | ···················· | ···················· |
| H | 8795 | hCoV-19/USA/LA-CDC-1284/2020\|EPI_ISL_535325\|2020-0 | ···················· | ···················· | ····················· | ···················· | ···················· | ···················· |
| H | 8796 | hCoV-19/USA/LA-CDC-1285/2020\|EPI_ISL_535326\|2020-0 | ···················· | ···················· | ····················· | ···················· | ···················· | ···················· |
| H | 8797 | hCoV-19/USA/LA-CDC-1286/2020\|EPI_ISL_535327\|2020-0 | ···················· | ···················· | ····················· | ···················· | ···················· | ···················· |
| H | 8798 | hCoV-19/USA/LA-CDC-1287/2020\|EPI_ISL_535328\|2020-0 | ···················· | ···················· | ····················· | ···················· | ···················· | ···················· |
| H | 8799 | hCoV-19/USA/LA-CDC-1288/2020\|EPI_ISL_535329\|2020-0 | ···················· | ···················· | ····················· | ···················· | ···················· | ···················· |
| H | 8800 | hCoV-19/USA/LA-CDC-1289/2020\|EPI_ISL_535330\|2020-0 | ···················· | ···················· | ····················· | ···················· | ···················· | ···················· |
| H | 8801 | hCoV-19/USA/LA-CDC-1290/2020\|EPI_ISL_535331\|2020-0 | ···················· | ···················· | ····················· | ···················· | ···················· | ···················· |
| H | 8802 | hCoV-19/USA/LA-CDC-1291/2020\|EPI_ISL_535332\|2020-0 | ···················· | ···················· | ····················· | ···················· | ···················· | ···················· |
| H | 8803 | hCoV-19/USA/LA-CDC-1292/2020\|EPI_ISL_535333\|2020-0 | ···················· | ···················· | ····················· | ···················· | ···················· | ···················· |
| H | 8804 | hCoV-19/USA/LA-CDC-1293/2020\|EPI_ISL_535334\|2020-0 | ···················· | ···················· | ····················· | ···················· | ···················· | ···················· |
| H | 8805 | hCoV-19/USA/LA-CDC-1294/2020\|EPI_ISL_535335\|2020-0 | ···················· | ···················· | ····················· | ···················· | ···················· | ···················· |
| H | 8806 | hCoV-19/USA/LA-CDC-1295/2020\|EPI_ISL_535336\|2020-0 | ···················· | ···················· | ····················· | ···················· | ···················· | ···················· |
| H | 8807 | hCoV-19/USA/LA-CDC-1296/2020\|EPI_ISL_535337\|2020-0 | ···················· | ···················· | ····················· | ···················· | ···················· | ···················· |
| H | 8808 | hCoV-19/USA/LA-CDC-1297/2020\|EPI_ISL_535338\|2020-0 | ···················· | ···················· | ····················· | ···················· | ···················· | ···················· |
| H | 8809 | hCoV-19/USA/LA-CDC-1298/2020\|EPI_ISL_535339\|2020-0 | ···················· | ···················· | ····················· | ···················· | ···················· | ···················· |
| H | 8810 | hCoV-19/USA/LA-CDC-1299/2020\|EPI_ISL_535340\|2020-0 | ···················· | ···················· | ····················· | ···················· | ···················· | ···················· |
| H | 8811 | hCoV-19/USA/LA-CDC-1905/2020\|EPI_ISL_535341\|2020-0 | ···················· | ···················· | ····················· | ···················· | ···················· | ···················· |
| H | 8812 | hCoV-19/USA/LA-CDC-1915/2020\|EPI_ISL_535342\|2020-0 | ···················· | ···················· | ····················· | ···················· | ···················· | ···················· |
| H | 8813 | hCoV-19/USA/LA-CDC-1916/2020\|EPI_ISL_535343\|2020-0 | ···················· | ···················· | ····················· | ···················· | ···················· | ···················· |
| H | 8814 | hCoV-19/USA/LA-CDC-1918/2020\|EPI_ISL_535344\|2020-0 | ···················· | ···················· | ····················· | ···················· | ···················· | ···················· |
| H | 8815 | hCoV-19/USA/LA-CDC-1932/2020\|EPI_ISL_535345\|2020-0 | ···················· | ···················· | ····················· | ···················· | ···················· | ···················· |
| H | 8816 | hCoV-19/USA/LA-CDC-2043/2020\|EPI_ISL_535346\|2020-0 | ···················· | ···················· | ····················· | ···················· | ···················· | ···················· |
| H | 8817 | hCoV-19/USA/LA-CDC-2103/2020\|EPI_ISL_535347\|2020-0 | ···················· | ···················· | ····················· | ···················· | ···················· | ···················· |
| H | 8818 | hCoV-19/USA/LA-CDC-2104/2020\|EPI_ISL_535348\|2020-0 | ···················· | ···················· | ····················· | ···················· | ···················· | ···················· |
| H | 8819 | hCoV-19/USA/LA-CDC-2111/2020\|EPI_ISL_535349\|2020-0 | ···················· | ···················· | ····················· | ···················· | ···················· | ···················· |
| H | 8820 | hCoV-19/USA/LA-CDC-2112/2020\|EPI_ISL_535350\|2020-0 | ···················· | ···················· | ····················· | ···················· | ···················· | ···················· |
| H | 8821 | hCoV-19/USA/LA-CDC-2119/2020\|EPI_ISL_535351\|2020-0 | ···················· | ···················· | ····················· | ···················· | ···················· | ···················· |
| H | 8822 | hCoV-19/USA/LA-CDC-2122/2020\|EPI_ISL_535352\|2020-0 | ···················· | ···················· | ····················· | ···················· | ···················· | ···················· |
| H | 8823 | hCoV-19/USA/LA-CDC-2145/2020\|EPI_ISL_535353\|2020-0 | ···················· | ···················· | ····················· | ···················· | ···················· | ···················· |
| H | 8824 | hCoV-19/USA/LA-CDC-2147/2020\|EPI_ISL_535354\|2020-0 | ···················· | ···················· | ····················· | ···················· | ···················· | ···················· |
| H | 8825 | hCoV-19/USA/LA-CDC-2148/2020\|EPI_ISL_535355\|2020-0 | ···················· | ···················· | ····················· | ···················· | ···················· | ···················· |
| H | 8826 | hCoV-19/USA/LA-CDC-2149/2020\|EPI_ISL_535356\|2020-0 | ···················· | ···················· | ····················· | ···················· | ···················· | ···················· |
| H | 8827 | hCoV-19/USA/LA-CDC-2153/2020\|EPI_ISL_535357\|2020-0 | ···················· | ···················· | ····················· | ···················· | ···················· | ···················· |
| H | 8828 | hCoV-19/USA/LA-CDC-2155/2020\|EPI_ISL_535358\|2020-0 | ···················· | ···················· | ····················· | ···················· | ···················· | ···················· |

| | | | | | | | | |
|---|---|---|---|---|---|---|---|---|
| H | 8829 | hCoV-19/USA/RI-CDC-6616/2020\|EPI_ISL_535359\|2020-0 | ···················· | ···················· | ···················· | ···················· | ···················· | ···················· |
| H | 8830 | hCoV-19/USA/RI-CDC-6619/2020\|EPI_ISL_535360\|2020-0 | ···················· | ···················· | ···················· | ···················· | ···················· | ···················· |
| H | 8831 | hCoV-19/USA/OK-ADDL-02/2020\|EPI_ISL_535361\|2020-05 | ···················· | ···················· | ···················· | ···················· | ···················· | ···················· |
| H | 8832 | hCoV-19/USA/OK-ADDL-03/2020\|EPI_ISL_535362\|2020-05 | ···················· | ···················· | ···················· | ···················· | ···················· | ···················· |
| H | 8833 | hCoV-19/USA/OK-ADDL-04/2020\|EPI_ISL_535363\|2020-05 | ···················· | ···················· | ···················· | ···················· | ···················· | ···················· |
| H | 8834 | hCoV-19/USA/OK-ADDL-01/2020\|EPI_ISL_535364\|2020-04 | ···················· | ·A·················· | ···················· | ···················· | ···················· | ···················· |
| H | 8835 | hCoV-19/USA/WI-UW-1228/2020\|EPI_ISL_536601\|2020-08 | ···················· | ···················· | ···················· | ···················· | ···················· | ···················· |
| H | 8836 | hCoV-19/USA/WI-UW-1229/2020\|EPI_ISL_536602\|2020-08 | ···················· | ···················· | ···················· | ···················· | ···················· | ···················· |
| H | 8837 | hCoV-19/USA/WI-UW-1230/2020\|EPI_ISL_536603\|2020-08 | ···················· | ···················· | ···················· | ···················· | ···················· | ···················· |
| H | 8838 | hCoV-19/USA/WI-UW-1253/2020\|EPI_ISL_536626\|2020-08 | ···················· | ···················· | ···················· | ···················· | ···················· | ···················· |
| H | 8839 | hCoV-19/USA/WI-UW-1254/2020\|EPI_ISL_536627\|2020-08 | ···················· | ···················· | ···················· | ···················· | ···················· | ···················· |
| H | 8840 | hCoV-19/USA/WI-UW-1255/2020\|EPI_ISL_536628\|2020-08 | ···················· | ···················· | ···················· | ···················· | ···················· | ···················· |
| H | 8841 | hCoV-19/USA/WI-UW-1256/2020\|EPI_ISL_536629\|2020-08 | ···················· | ···················· | ···················· | ···················· | ···················· | ···················· |
| H | 8842 | hCoV-19/USA/WI-UW-1257/2020\|EPI_ISL_536630\|2020-08 | ···················· | ·A·················· | ···················· | ···················· | ···················· | ···················· |
| H | 8843 | hCoV-19/USA/WI-UW-1258/2020\|EPI_ISL_536631\|2020-08 | ···················· | ···················· | ···················· | ···················· | ···················· | ···················· |
| H | 8844 | hCoV-19/USA/WI-UW-1259/2020\|EPI_ISL_536632\|2020-08 | ···················· | ···················· | ···················· | ···················· | ···················· | ···················· |
| H | 8845 | hCoV-19/USA/WI-UW-1260/2020\|EPI_ISL_536633\|2020-08 | ···················· | ·A·················· | ···················· | ···················· | ···················· | ···················· |
| H | 8846 | hCoV-19/USA/WI-UW-1261/2020\|EPI_ISL_536634\|2020-08 | ···················· | ···················· | ···················· | ···················· | ···················· | ···················· |
| H | 8847 | hCoV-19/USA/WI-UW-1262/2020\|EPI_ISL_536635\|2020-08 | ···················· | ···················· | ···················· | ···················· | ···················· | ···················· |
| H | 8848 | hCoV-19/USA/WI-UW-1263/2020\|EPI_ISL_536636\|2020-08 | ···················· | ···················· | ···················· | ···················· | ···················· | ···················· |
| H | 8849 | hCoV-19/USA/WI-UW-1264/2020\|EPI_ISL_536637\|2020-08 | ···················· | ·A·················· | ···················· | ···················· | ···················· | ···················· |
| H | 8850 | hCoV-19/USA/WI-UW-1266/2020\|EPI_ISL_536638\|2020-08 | ···················· | ···················· | ···················· | ···················· | ···················· | ···················· |
| H | 8851 | hCoV-19/USA/WI-UW-1267/2020\|EPI_ISL_536639\|2020-08 | ···················· | ···················· | ···················· | ···················· | ···················· | ···················· |
| H | 8852 | hCoV-19/USA/WI-UW-1268/2020\|EPI_ISL_536640\|2020-08 | ···················· | ···················· | ···················· | ···················· | ···················· | ···················· |
| H | 8853 | hCoV-19/USA/WI-UW-1269/2020\|EPI_ISL_536641\|2020-08 | ···················· | ···················· | ···················· | ···················· | ···················· | ···················· |
| H | 8854 | hCoV-19/USA/WI-UW-1270/2020\|EPI_ISL_536642\|2020-08 | ···················· | ···················· | ···················· | ···················· | ···················· | ···················· |
| H | 8855 | hCoV-19/USA/WI-UW-1271/2020\|EPI_ISL_536643\|2020-08 | ···················· | ···················· | ···················· | ···················· | ···················· | ···················· |
| H | 8856 | hCoV-19/USA/WI-UW-1272/2020\|EPI_ISL_536644\|2020-08 | ···················· | ···················· | ···················· | ···················· | ···················· | ···················· |
| H | 8857 | hCoV-19/USA/WI-UW-1273/2020\|EPI_ISL_536645\|2020-08 | ···················· | ···················· | ···················· | ···················· | ···················· | ···················· |
| H | 8858 | hCoV-19/USA/WI-UW-1274/2020\|EPI_ISL_536646\|2020-08 | ···················· | ···················· | ···················· | ···················· | ···················· | ···················· |
| H | 8859 | hCoV-19/USA/WI-UW-1275/2020\|EPI_ISL_536647\|2020-08 | ···················· | ···················· | ···················· | ···················· | ···················· | ···················· |
| H | 8860 | hCoV-19/USA/WI-UW-1276/2020\|EPI_ISL_536648\|2020-08 | ···················· | ···················· | ···················· | ···················· | ···················· | ···················· |
| H | 8861 | hCoV-19/USA/WI-UW-1277/2020\|EPI_ISL_536649\|2020-08 | ···················· | ···················· | ···················· | ···················· | ···················· | ···················· |
| H | 8862 | hCoV-19/USA/WI-UW-1278/2020\|EPI_ISL_536650\|2020-08 | ···················· | ···················· | ···················· | ···················· | ···················· | ···················· |
| H | 8863 | hCoV-19/USA/WI-UW-1279/2020\|EPI_ISL_536651\|2020-08 | ···················· | ·A·················· | ···················· | ···················· | ···················· | ···················· |
| H | 8864 | hCoV-19/USA/WI-UW-1280/2020\|EPI_ISL_536652\|2020-08 | ···················· | ···················· | ···················· | ···················· | ···················· | ···················· |
| H | 8865 | hCoV-19/USA/WI-UW-1281/2020\|EPI_ISL_536653\|2020-08 | ···················· | ···················· | ···················· | ···················· | ···················· | ···················· |
| H | 8866 | hCoV-19/USA/WI-UW-1282/2020\|EPI_ISL_536654\|2020-08 | ···················· | ···················· | ···················· | ···················· | ···················· | ···················· |
| H | 8867 | hCoV-19/USA/WI-UW-1283/2020\|EPI_ISL_536655\|2020-08 | ···················· | ·A·················· | ···················· | ···················· | ···················· | ···················· |
| H | 8868 | hCoV-19/USA/WI-UW-1284/2020\|EPI_ISL_536656\|2020-09 | ···················· | ···················· | ···················· | ···················· | ···················· | ···················· |
| H | 8869 | hCoV-19/USA/WI-UW-1285/2020\|EPI_ISL_536657\|2020-09 | ···················· | ···················· | ···················· | ···················· | ···················· | ···················· |
| H | 8870 | hCoV-19/USA/WI-UW-1286/2020\|EPI_ISL_536658\|2020-08 | ···················· | ···················· | ···················· | ···················· | ···················· | ···················· |
| H | 8871 | hCoV-19/USA/WI-UW-1288/2020\|EPI_ISL_536660\|2020-09 | ···················· | ·A·················· | ···················· | ···················· | ···················· | ···················· |
| H | 8872 | hCoV-19/USA/NY-UW-1289/2020\|EPI_ISL_536661\|2020-09 | ···················· | ···················· | ···················· | ···················· | ···················· | ···················· |
| H | 8873 | hCoV-19/USA/WI-UW-1290/2020\|EPI_ISL_536662\|2020-09 | ···················· | ···················· | ···················· | ···················· | ···················· | ···················· |
| H | 8874 | hCoV-19/USA/WI-UW-1291/2020\|EPI_ISL_536663\|2020-08 | ···················· | ···················· | ···················· | ···················· | ···················· | ···················· |
| H | 8875 | hCoV-19/USA/WI-UW-1292/2020\|EPI_ISL_536664\|2020-09 | ···················· | ···················· | ···················· | ···················· | ···················· | ···················· |
| H | 8876 | hCoV-19/USA/WI-UW-1293/2020\|EPI_ISL_536665\|2020-09 | ···················· | ···················· | ···················· | ···················· | ···················· | ···················· |
| H | 8877 | hCoV-19/USA/WI-UW-1294/2020\|EPI_ISL_536666\|2020-09 | ···················· | ···················· | ···················· | ···················· | ···················· | ···················· |
| H | 8878 | hCoV-19/USA/WI-UW-1295/2020\|EPI_ISL_536667\|2020-09 | ···················· | ··C················· | ···················· | ···················· | ···················· | ···················· |
| H | 8879 | hCoV-19/USA/CA-UW-1296/2020\|EPI_ISL_536668\|2020-09 | ···················· | ···················· | ···················· | ···················· | ···················· | ···················· |
| H | 8880 | hCoV-19/USA/WI-UW-1297/2020\|EPI_ISL_536669\|2020-08 | ···················· | ···················· | ···················· | ···················· | ···················· | ···················· |
| H | 8881 | hCoV-19/USA/WI-UW-1298/2020\|EPI_ISL_536670\|2020-08 | ···················· | ···················· | ···················· | ···················· | ···················· | ···················· |
| H | 8882 | hCoV-19/USA/WI-UW-1299/2020\|EPI_ISL_536671\|2020-08 | ···················· | ···················· | ···················· | ···················· | ···················· | ···················· |
| H | 8883 | hCoV-19/USA/WI-UW-1301/2020\|EPI_ISL_536673\|2020-08 | ···················· | ···················· | ···················· | ···················· | ···················· | ···················· |
| H | 8884 | hCoV-19/USA/WI-UW-1302/2020\|EPI_ISL_536674\|2020-09 | ···················· | ···················· | ···················· | ···················· | ···················· | ···················· |
| H | 8885 | hCoV-19/USA/WI-UW-1303/2020\|EPI_ISL_536675\|2020-08 | ···················· | ·A·················· | ···················· | ···················· | ···················· | ···················· |
| H | 8886 | hCoV-19/USA/WI-UW-1304/2020\|EPI_ISL_536676\|2020-09 | ···················· | ···················· | ···················· | ···················· | ···················· | ···················· |
| H | 8887 | hCoV-19/USA/WI-UW-1305/2020\|EPI_ISL_536677\|2020-08 | ···················· | ···················· | ···················· | ···················· | ···················· | ···················· |
| H | 8888 | hCoV-19/USA/CA-UW-1306/2020\|EPI_ISL_536678\|2020-09 | ···················· | ···················· | ···················· | ···················· | ···················· | ···················· |
| H | 8895 | hCoV-19/USA/WI-UW-1314/2020\|EPI_ISL_536686\|2020-08 | ···················· | ···················· | ···················· | ···················· | ···················· | ···················· |
| H | 8900 | hCoV-19/USA/WI-UW-1326/2020\|EPI_ISL_536697\|2020-08 | ···················· | ···················· | ···················· | ···················· | ···················· | ···················· |
| H | 8901 | hCoV-19/USA/WI-UW-1327/2020\|EPI_ISL_536698\|2020-08 | ···················· | ···················· | ···················· | ···················· | ···················· | ···················· |
| H | 8902 | hCoV-19/USA/WI-UW-1328/2020\|EPI_ISL_536699\|2020-08 | ···················· | ···················· | ···················· | ···················· | ···················· | ···················· |
| H | 8903 | hCoV-19/Czech Republic/NRL-1753/2020\|EPI_ISL_54113 | ···················· | ···················· | ···················· | ···················· | ···················· | ···················· |
| H | 8904 | hCoV-19/USA/FL-BPHL-1259/2020\|EPI_ISL_541144\|2020- | ···················· | ···················· | ···················· | ···················· | ···················· | ···················· |
| H | 8907 | hCoV-19/USA/FL-BPHL-1249/2020\|EPI_ISL_541155\|2020- | ···················· | ···················· | ···················· | ···················· | ···················· | ···················· |
| H | 8908 | hCoV-19/USA/FL-BPHL-1234/2020\|EPI_ISL_541158\|2020- | ···················· | ···················· | ···················· | ···················· | ···················· | ···················· |
| H | 8909 | hCoV-19/USA/FL-BPHL-1241/2020\|EPI_ISL_541164\|2020- | ···················· | ···················· | ···················· | ···················· | ···················· | ···················· |
| H | 8910 | hCoV-19/USA/FL-BPHL-1261/2020\|EPI_ISL_541168\|2020- | ···················· | ···················· | ···················· | ···················· | ···················· | ···················· |
| H | 8913 | hCoV-19/USA/FL-BPHL-1202/2020\|EPI_ISL_541175\|2020- | ···················· | ···················· | ···················· | ···················· | ···················· | ···················· |
| H | 8914 | hCoV-19/USA/FL-BPHL-1203/2020\|EPI_ISL_541176\|2020- | ···················· | ···················· | ···················· | ···················· | ···················· | ···················· |
| H | 8915 | hCoV-19/USA/FL-BPHL-1205/2020\|EPI_ISL_541178\|2020- | ···················· | ···················· | ···················· | ···················· | ···················· | ···················· |
| H | 8916 | hCoV-19/USA/FL-BPHL-1206/2020\|EPI_ISL_541179\|2020- | ···················· | ···················· | ···················· | ···················· | ···················· | ···················· |
| H | 8918 | hCoV-19/USA/FL-BPHL-1208/2020\|EPI_ISL_541181\|2020- | ···················· | ···················· | ···················· | ···················· | ···················· | ···················· |
| H | 8919 | hCoV-19/USA/FL-BPHL-1209/2020\|EPI_ISL_541182\|2020- | ···················· | ····A··············· | ···················· | ···················· | ···················· | ···················· |
| H | 8920 | hCoV-19/USA/FL-BPHL-1212/2020\|EPI_ISL_541185\|2020- | ···················· | ···················· | ···················· | ···················· | ···················· | ···················· |
| H | 8921 | hCoV-19/USA/FL-BPHL-1213/2020\|EPI_ISL_541186\|2020- | ···················· | ···················· | ···················· | ···················· | ···················· | ···················· |
| H | 8922 | hCoV-19/USA/FL-BPHL-1214/2020\|EPI_ISL_541187\|2020- | ···················· | ···················· | ···················· | ···················· | ···················· | ···················· |
| H | 8923 | hCoV-19/USA/FL-BPHL-1215/2020\|EPI_ISL_541188\|2020- | ···················· | ···················· | ···················· | ···················· | ···················· | ···················· |

| | | | | | | | | |
|---|---|---|---|---|---|---|---|---|
| H | 8924 | hCoV-19/USA/FL-BPHL-1216/2020\|EPI_ISL_541189\|2020- | ···················· | ···················· | ···················· | ···················· | ···················· | ···················· |
| H | 8925 | hCoV-19/USA/FL-BPHL-1218/2020\|EPI_ISL_541191\|2020- | ···················· | ···················· | ···················· | ···················· | ···················· | ···················· |
| H | 8926 | hCoV-19/USA/FL-BPHL-1219/2020\|EPI_ISL_541192\|2020- | ···················· | ···················· | ···················· | ···················· | ···················· | ···················· |
| H | 8928 | hCoV-19/USA/FL-BPHL-1222/2020\|EPI_ISL_541195\|2020- | ···················· | ···················· | ···················· | ···················· | ···················· | ···················· |
| H | 8929 | hCoV-19/USA/FL-BPHL-1223/2020\|EPI_ISL_541196\|2020- | ···················· | ···················· | ···················· | ···················· | ···················· | ···················· |
| H | 8931 | hCoV-19/USA/FL-BPHL-1226/2020\|EPI_ISL_541199\|2020- | ···················· | ···················· | ···················· | ···················· | ···················· | ···················· |
| H | 8932 | hCoV-19/USA/TX-HMH-1002/2020\|EPI_ISL_542487\|2020-0 | ···················· | ···················· | ···················· | ···················· | ···················· | ···················· |
| H | 8933 | hCoV-19/USA/TX-HMH-1003/2020\|EPI_ISL_542488\|2020-0 | ···················· | ···················· | ···················· | ···················· | ···················· | ···················· |
| H | 8934 | hCoV-19/USA/TX-HMH-1007/2020\|EPI_ISL_542491\|2020-0 | ···················· | ···················· | ···················· | ···················· | ···················· | ···················· |
| H | 8935 | hCoV-19/USA/TX-HMH-1011/2020\|EPI_ISL_542492\|2020-0 | ···················· | ···················· | ···················· | ···················· | ···················· | ···················· |
| H | 8936 | hCoV-19/USA/TX-HMH-1012/2020\|EPI_ISL_542493\|2020-0 | ···················· | ···················· | ···················· | ···················· | ···················· | ···················· |
| H | 8937 | hCoV-19/USA/TX-HMH-1013/2020\|EPI_ISL_542494\|2020-0 | ···················· | ···················· | ···················· | ···················· | ···················· | ···················· |
| H | 8938 | hCoV-19/USA/TX-HMH-1016/2020\|EPI_ISL_542495\|2020-0 | ···················· | ···················· | ···················· | ···················· | ···················· | ···················· |
| H | 8939 | hCoV-19/USA/TX-HMH-1020/2020\|EPI_ISL_542496\|2020-0 | ···················· | ···················· | ···················· | ···················· | ···················· | ···················· |
| H | 8940 | hCoV-19/USA/TX-HMH-1022/2020\|EPI_ISL_542497\|2020-0 | ···················· | ···················· | ···················· | ···················· | ···················· | ···················· |
| H | 8941 | hCoV-19/USA/TX-HMH-1024/2020\|EPI_ISL_542498\|2020-0 | ···················· | ···················· | ···················· | ···················· | ···················· | ···················· |
| H | 8942 | hCoV-19/USA/TX-HMH-1235/2020\|EPI_ISL_542600\|2020-0 | ···················· | ···················· | ···················· | ···················· | ···················· | ···················· |
| H | 8943 | hCoV-19/USA/TX-HMH-1236/2020\|EPI_ISL_542601\|2020-0 | ···················· | ···················· | ···················· | ···················· | ···················· | ···················· |
| H | 8944 | hCoV-19/USA/TX-HMH-1237/2020\|EPI_ISL_542602\|2020-0 | ···················· | ···················· | ···················· | ···················· | ···················· | ···················· |
| H | 8945 | hCoV-19/USA/TX-HMH-1238/2020\|EPI_ISL_542603\|2020-0 | ···················· | ···················· | ···················· | ···················· | ···················· | ···················· |
| H | 8946 | hCoV-19/USA/TX-HMH-1240/2020\|EPI_ISL_542604\|2020-0 | ···················· | ···················· | ···················· | ···················· | ···················· | ···················· |
| H | 8947 | hCoV-19/USA/TX-HMH-1241/2020\|EPI_ISL_542605\|2020-0 | ···················· | ···················· | ···················· | ···················· | ···················· | ···················· |
| H | 8948 | hCoV-19/USA/TX-HMH-1242/2020\|EPI_ISL_542606\|2020-0 | ···················· | ···················· | ···················· | ···················· | ···················· | ···················· |
| H | 8949 | hCoV-19/USA/TX-HMH-1244/2020\|EPI_ISL_542607\|2020-0 | ···················· | ···················· | ···················· | ···················· | ···················· | ···················· |
| H | 8950 | hCoV-19/USA/TX-HMH-1247/2020\|EPI_ISL_542609\|2020-0 | ···················· | ···················· | ···················· | ···················· | ···················· | ···················· |
| H | 8951 | hCoV-19/USA/TX-HMH-1248/2020\|EPI_ISL_542610\|2020-0 | ···················· | ···················· | ···················· | ···················· | ···················· | ···················· |
| H | 8952 | hCoV-19/USA/TX-HMH-1249/2020\|EPI_ISL_542611\|2020-0 | ···················· | ···················· | ···················· | ···················· | ···················· | ···················· |
| H | 8953 | hCoV-19/USA/TX-HMH-1250/2020\|EPI_ISL_542612\|2020-0 | ···················· | ···················· | ···················· | ···················· | ···················· | ···················· |
| H | 8954 | hCoV-19/USA/TX-HMH-1252/2020\|EPI_ISL_542613\|2020-0 | ···················· | ···················· | ···················· | ···················· | ···················· | ···················· |
| H | 8955 | hCoV-19/USA/TX-HMH-1254/2020\|EPI_ISL_542614\|2020-0 | ···················· | ···················· | ···················· | ···················· | ···················· | ···················· |
| H | 8956 | hCoV-19/USA/TX-HMH-1256/2020\|EPI_ISL_542615\|2020-0 | ···················· | ···················· | ···················· | ···················· | ···················· | ···················· |
| H | 8958 | hCoV-19/USA/TX-HMH-1263/2020\|EPI_ISL_542617\|2020-0 | ···················· | ···················· | ···················· | ···················· | ···················· | ···················· |
| H | 8959 | hCoV-19/USA/TX-HMH-1264/2020\|EPI_ISL_542618\|2020-0 | ···················· | ···················· | ···················· | ···················· | ···················· | ···················· |
| H | 8960 | hCoV-19/USA/TX-HMH-1270/2020\|EPI_ISL_542620\|2020-0 | ···················· | ···················· | ···················· | ···················· | ···················· | ···················· |
| H | 8961 | hCoV-19/England/QEUH-96F3A3/2020\|EPI_ISL_530521\|20 | ···················· | ···················· | ···················· | ···················· | ···················· | ···················· |
| H | 8962 | hCoV-19/Scotland/QEUH-96E9B1/2020\|EPI_ISL_530523\|2 | ···················· | ···················· | ···················· | ···················· | ···················· | ···················· |
| H | 8963 | hCoV-19/Scotland/QEUH-96E8F0/2020\|EPI_ISL_530528\|2 | ···················· | ···················· | ···················· | ···················· | ···················· | ···················· |
| H | 8964 | hCoV-19/England/QEUH-96F2C4/2020\|EPI_ISL_530530\|20 | ···················· | ···················· | ···················· | ···················· | ···················· | ···················· |
| H | 8965 | hCoV-19/England/QEUH-96DE86/2020\|EPI_ISL_530531\|20 | ···················· | ···················· | ···················· | ···················· | ···················· | ···················· |
| H | 8966 | hCoV-19/England/QEUH-96F8B3/2020\|EPI_ISL_530532\|20 | ···················· | ···················· | ···················· | ···················· | ···················· | ···················· |
| H | 8967 | hCoV-19/Scotland/QEUH-96E580/2020\|EPI_ISL_530533\|2 | ···················· | ···················· | ···················· | ···················· | ···················· | ···················· |
| H | 8968 | hCoV-19/England/QEUH-96E6BA/2020\|EPI_ISL_530534\|20 | ···················· | ···················· | ···················· | ···················· | ···················· | ···················· |
| H | 8969 | hCoV-19/England/QEUH-96E9C0/2020\|EPI_ISL_530536\|20 | ···················· | ···················· | ···················· | ···················· | ···················· | ···················· |
| H | 8970 | hCoV-19/England/QEUH-96F23D/2020\|EPI_ISL_530537\|20 | ···················· | ···················· | ···················· | ···················· | ···················· | ···················· |
| H | 8971 | hCoV-19/England/QEUH-96E632/2020\|EPI_ISL_530538\|20 | ···················· | ···················· | ···················· | ···················· | ···················· | ···················· |
| H | 8972 | hCoV-19/England/QEUH-96E562/2020\|EPI_ISL_530540\|20 | ···················· | ···················· | ···················· | ···················· | ·····T·············· | ···················· |
| H | 8973 | hCoV-19/England/QEUH-96E5DB/2020\|EPI_ISL_530541\|20 | ···················· | ·A·················· | ···················· | ···················· | ···················· | ···················· |
| H | 8974 | hCoV-19/Scotland/QEUH-96E41A/2020\|EPI_ISL_530542\|2 | ···················· | ···················· | ···················· | ···················· | ···················· | ···················· |
| H | 8975 | hCoV-19/England/QEUH-96F491/2020\|EPI_ISL_530543\|20 | ···················· | ···················· | ···················· | ···················· | ···················· | ···················· |
| H | 8976 | hCoV-19/England/QEUH-96E720/2020\|EPI_ISL_530544\|20 | ···················· | ···················· | ···················· | ···················· | ···················· | ···················· |
| H | 8977 | hCoV-19/England/QEUH-96F1A9/2020\|EPI_ISL_530545\|20 | ···················· | ···················· | ···················· | ···················· | ···················· | ···················· |
| H | 8978 | hCoV-19/England/QEUH-96EA18/2020\|EPI_ISL_530546\|20 | ···················· | ···················· | ···················· | ···················· | ···················· | ···················· |
| H | 8979 | hCoV-19/England/QEUH-96E6F6/2020\|EPI_ISL_530547\|20 | ···················· | ···················· | ···················· | ···················· | ···················· | ···················· |
| H | 8980 | hCoV-19/England/QEUH-96F22E/2020\|EPI_ISL_530548\|20 | ···················· | ···················· | ···················· | ···················· | ···················· | ···················· |
| H | 8981 | hCoV-19/England/QEUH-96F4FB/2020\|EPI_ISL_530549\|20 | ···················· | ···················· | ···················· | ···················· | ···················· | ···················· |
| H | 8982 | hCoV-19/England/QEUH-96EF55/2020\|EPI_ISL_530550\|20 | ···················· | ···················· | ···················· | ···················· | ···················· | ···················· |
| H | 8983 | hCoV-19/England/QEUH-96E9FD/2020\|EPI_ISL_530551\|20 | ···················· | ···················· | ···················· | ···················· | ···················· | ···················· |
| H | 8984 | hCoV-19/England/QEUH-96E9DF/2020\|EPI_ISL_530552\|20 | ···················· | ···················· | ···················· | ···················· | ···················· | ···················· |
| H | 8985 | hCoV-19/Scotland/QEUH-96EBF7/2020\|EPI_ISL_530553\|2 | ···················· | ·A·················· | ···················· | ···················· | ···················· | ··A················· |
| H | 8986 | hCoV-19/England/QEUH-96E2B6/2020\|EPI_ISL_530554\|20 | ···················· | ···················· | ···················· | ···················· | ···················· | ···················· |
| H | 8987 | hCoV-19/England/QEUH-96F279/2020\|EPI_ISL_530555\|20 | ···················· | ···················· | ···················· | ···················· | ···················· | ···················· |
| H | 8988 | hCoV-19/England/QEUH-96DF83/2020\|EPI_ISL_530556\|20 | ···················· | ···················· | ···················· | ···················· | ···················· | ···················· |
| H | 8989 | hCoV-19/Northern Ireland/QEUH-96F006/2020\|EPI_ISL_ | ···················· | ···················· | ···················· | ···················· | ···················· | ···················· |
| H | 8990 | hCoV-19/England/QEUH-96E201/2020\|EPI_ISL_530559\|20 | ···················· | ···················· | ···················· | ···················· | ···················· | ···················· |
| H | 8991 | hCoV-19/Northern Ireland/QEUH-96ED2E/2020\|EPI_ISL_ | ···················· | ···················· | ···················· | ···················· | ···················· | ···················· |
| H | 8992 | hCoV-19/Northern Ireland/QEUH-96EA36/2020\|EPI_ISL_ | ···················· | ···················· | ···················· | ···················· | ···················· | ···················· |
| H | 8993 | hCoV-19/Scotland/QEUH-96E18C/2020\|EPI_ISL_530562\|2 | ···················· | ···················· | ···················· | ···················· | ···················· | ···················· |
| H | 8994 | hCoV-19/Northern Ireland/QEUH-96EC30/2020\|EPI_ISL_ | ···················· | ···················· | ···················· | ···················· | ···················· | ···················· |
| H | 8995 | hCoV-19/England/QEUH-96F6D7/2020\|EPI_ISL_530564\|20 | ···················· | ···················· | ···················· | ···················· | ···················· | ···················· |
| H | 8996 | hCoV-19/England/QEUH-96E0E9/2020\|EPI_ISL_530565\|20 | ···················· | ···················· | ···················· | ···················· | ···················· | ···················· |
| H | 8997 | hCoV-19/Northern Ireland/QEUH-96EA81/2020\|EPI_ISL_ | ···················· | ···················· | ···················· | ···················· | ···················· | ···················· |
| H | 8998 | hCoV-19/England/QEUH-96F3C1/2020\|EPI_ISL_530567\|20 | ···················· | ···················· | ···················· | ···················· | ···················· | ···················· |
| H | 8999 | hCoV-19/England/QEUH-96F473/2020\|EPI_ISL_530568\|20 | ···················· | ···················· | ···················· | ···················· | ···················· | ···················· |
| H | 9000 | hCoV-19/England/QEUH-96E80F/2020\|EPI_ISL_530570\|20 | ···················· | ···················· | ···················· | ···················· | ···················· | ···················· |
| H | 9001 | hCoV-19/England/QEUH-96E571/2020\|EPI_ISL_530571\|20 | ···················· | ···················· | ···················· | ···················· | ···················· | ···················· |
| H | 9002 | hCoV-19/England/QEUH-96F437/2020\|EPI_ISL_530572\|20 | ···················· | ···················· | ···················· | ···················· | ···················· | ···················· |
| H | 9003 | hCoV-19/England/QEUH-96E40B/2020\|EPI_ISL_530573\|20 | ···················· | ···················· | ···················· | ···················· | ···················· | ···················· |
| H | 9004 | hCoV-19/England/QEUH-96F25B/2020\|EPI_ISL_530574\|20 | ···················· | ···················· | ···················· | ···················· | ···················· | ···················· |
| H | 9005 | hCoV-19/England/QEUH-96F376/2020\|EPI_ISL_530575\|20 | ···················· | ···················· | ···················· | ···················· | ···················· | ···················· |
| H | 9006 | hCoV-19/Northern Ireland/QEUH-96F09D/2020\|EPI_ISL_ | ···················· | ···················· | ···················· | ···················· | ···················· | ···················· |

```
H    9007 hCoV-19/England/QEUH-96F358/2020|EPI_ISL_530578|20    ....................  ....................  ....................  ....................  ....................  ....................
H    9008 hCoV-19/England/QEUH-96ED5B/2020|EPI_ISL_530580|20    ....................  ....................  ....................  ....................  ....................  ....................
H    9009 hCoV-19/England/QEUH-96E605/2020|EPI_ISL_530581|20    ....................  ....................  ....................  ....................  ....................  ....................
H    9010 hCoV-19/England/QEUH-96DEE0/2020|EPI_ISL_530582|20    ....................  ....................  ....................  ....................  ....................  ....................
H    9011 hCoV-19/England/QEUH-96E7B7/2020|EPI_ISL_530583|20    ....................  ....................  ....................  ....................  ....................  ....................
H    9012 hCoV-19/Northern Ireland/QEUH-96EC6D/2020|EPI_ISL_    ....................  ....................  ....................  ....................  ....................  ....................
H    9013 hCoV-19/England/QEUH-96E7D5/2020|EPI_ISL_530585|20    ....................  ....................  ....................  ....................  ....................  ....................
H    9014 hCoV-19/England/QEUH-96F33A/2020|EPI_ISL_530586|20    ....................  .A..................  ....................  ....................  ....................  ....................
H    9015 hCoV-19/England/QEUH-96E641/2020|EPI_ISL_530587|20    ....................  ....................  ....................  ....................  ....................  ....................
H    9016 hCoV-19/England/QEUH-96F16D/2020|EPI_ISL_530589|20    ....................  ....................  ....................  ....................  ....................  ....................
H    9017 hCoV-19/England/QEUH-96DF47/2020|EPI_ISL_530590|20    ....................  ....................  ....................  ....................  ....................  ....................
H    9018 hCoV-19/England/QEUH-96E711/2020|EPI_ISL_530591|20    ....................  ....................  ....................  ....................  ....................  ....................
H    9019 hCoV-19/England/QEUH-96E025/2020|EPI_ISL_530593|20    ....................  ....................  ....................  ....................  ....................  ....................
H    9020 hCoV-19/England/QEUH-96F4DD/2020|EPI_ISL_530594|20    ....................  ....................  ....................  ....................  ....................  ....................
H    9021 hCoV-19/England/QEUH-96F3FE/2020|EPI_ISL_530595|20    ....................  ....................  ....................  ....................  ....................  ....................
H    9022 hCoV-19/Scotland/QEUH-96E5CC/2020|EPI_ISL_530596|2    ....................  ....................  ....................  ....................  ....................  ....................
H    9023 hCoV-19/England/QEUH-96EB60/2020|EPI_ISL_530597|20    ....................  ....................  ....................  ....................  ....................  ....................
H    9024 hCoV-19/England/QEUH-96E27A/2020|EPI_ISL_530598|20    ....................  ....................  ....................  ....................  ....................  ....................
H    9025 hCoV-19/England/QEUH-96E22F/2020|EPI_ISL_530599|20    ....................  ....................  ....................  ....................  ....................  ....................
H    9026 hCoV-19/England/QEUH-9621F1/2020|EPI_ISL_531800|20    ....................  ....................  ....................  ....................  ....................  ....................
H    9027 hCoV-19/Scotland/QEUH-9631E1/2020|EPI_ISL_531801|2    ....................  ....................  ....................  ....................  ....................  ....................
H    9028 hCoV-19/Scotland/QEUH-962B3F/2020|EPI_ISL_531802|2    ....................  ....................  ....................  ....................  ....................  ....................
H    9029 hCoV-19/Northern Ireland/QEUH-962197/2020|EPI_ISL_    ....................  ....................  ....................  ....................  ....................  ....................
H    9030 hCoV-19/Northern Ireland/QEUH-9622FE/2020|EPI_ISL_    ....................  ....................  ....................  ....................  ....................  ....................
H    9031 hCoV-19/Scotland/QEUH-962BA8/2020|EPI_ISL_531805|2    ....................  ....................  ....................  ....................  ....................  ....................
H    9032 hCoV-19/England/QEUH-963776/2020|EPI_ISL_531806|20    ....................  ....................  ....................  ....................  ....................  ....................
H    9033 hCoV-19/Northern Ireland/QEUH-962337/2020|EPI_ISL_    ....................  ....................  ....................  ....................  ....................  ....................
H    9034 hCoV-19/England/QEUH-962F8E/2020|EPI_ISL_531852|20    ....................  ....................  ....................  ....................  ....................  ....................
H    9035 hCoV-19/England/QEUH-963503/2020|EPI_ISL_531853|20    ....................  ....................  ....................  ....................  ....................  ....................
H    9036 hCoV-19/England/QEUH-963697/2020|EPI_ISL_531854|20    ....................  ....................  ....................  ....................  ....................  ....................
H    9037 hCoV-19/Scotland/QEUH-961F9E/2020|EPI_ISL_531856|2    ....................  ....................  ....................  ....................  ....................  ....................
H    9038 hCoV-19/Scotland/QEUH-961F34/2020|EPI_ISL_531857|2    ....................  ....................  ....................  ....................  ....................  ....................
H    9039 hCoV-19/Scotland/QEUH-962AF6/2020|EPI_ISL_531858|2    ....................  ....................  ....................  ....................  ....................  ....................
H    9040 hCoV-19/England/QEUH-962A41/2020|EPI_ISL_531859|20    ....................  ....................  ....................  ....................  ....................  ....................
H    9041 hCoV-19/England/QEUH-96273B/2020|EPI_ISL_531860|20    ....................  ....................  ....................  ....................  ....................  ....................
H    9042 hCoV-19/England/QEUH-962D2A/2020|EPI_ISL_531861|20    ....................  ....................  ....................  ....................  ....................  ....................
H    9043 hCoV-19/Scotland/QEUH-963530/2020|EPI_ISL_531862|2    ....................  ....................  ....................  ....................  ....................  ....................
H    9044 hCoV-19/Scotland/QEUH-96355E/2020|EPI_ISL_531863|2    ....................  ....................  ....................  ....................  ....................  ....................
H    9045 hCoV-19/England/QEUH-96017B/2020|EPI_ISL_531864|20    ....................  ....................  ....................  ....................  ....................  ....................
H    9046 hCoV-19/Scotland/QEUH-95FE0F/2020|EPI_ISL_531865|2    ....................  ....................  ....................  ....................  ....................  ....................
H    9047 hCoV-19/Northern Ireland/QEUH-95FEB4/2020|EPI_ISL_    ....................  ....................  ....................  ....................  ....................  ....................
H    9048 hCoV-19/England/QEUH-95FE96/2020|EPI_ISL_531867|20    ....................  ....................  ....................  ....................  ....................  ....................
H    9049 hCoV-19/England/QEUH-9601C6/2020|EPI_ISL_531868|20    ....................  ....................  ....................  ....................  ....................  ....................
H    9050 hCoV-19/England/QEUH-95FF1B/2020|EPI_ISL_531869|20    ....................  .A..................  ....................  ....................  ....................  ....................
H    9051 hCoV-19/Northern Ireland/QEUH-95FFB1/2020|EPI_ISL_    ....................  ....................  ....................  ....................  ....................  ....................
H    9052 hCoV-19/England/QEUH-95FD99/2020|EPI_ISL_531871|20    ....................  ....................  ....................  ....................  ....................  ....................
H    9053 hCoV-19/Scotland/QEUH-9600F6/2020|EPI_ISL_531873|2    ....................  ....................  ....................  ....................  ....................  ....................
H    9054 hCoV-19/England/QEUH-95FD11/2020|EPI_ISL_531874|20    ....................  ....................  ....................  ....................  ....................  ....................
H    9055 hCoV-19/England/QEUH-95FD3F/2020|EPI_ISL_531875|20    ....................  .A.......A..........  ....................  ....................  ....................  ....................
H    9056 hCoV-19/England/QEUH-95FBAE/2020|EPI_ISL_531876|20    ....................  ....................  ....................  ....................  ....................  ....................
H    9057 hCoV-19/Scotland/QEUH-960445/2020|EPI_ISL_531877|2    ....................  ....................  ....................  ....................  ....................  ....................
H    9058 hCoV-19/England/QEUH-95FD7B/2020|EPI_ISL_531879|20    ....................  ....................  ....................  ....................  ....................  ....................
H    9059 hCoV-19/Scotland/QEUH-960014/2020|EPI_ISL_531880|2    ....................  ....................  ....................  ....................  ....................  ....................
H    9060 hCoV-19/Scotland/QEUH-95FFFD/2020|EPI_ISL_531881|2    ....................  ....................  ....................  ....................  ....................  ....................
H    9061 hCoV-19/England/QEUH-960111/2020|EPI_ISL_531882|20    ....................  ....................  ....................  ....................  ....................  ....................
H    9062 hCoV-19/England/QEUH-960348/2020|EPI_ISL_531883|20    ....................  .A..................  ....................  ....................  ....................  ....................
H    9063 hCoV-19/Scotland/QEUH-9600D8/2020|EPI_ISL_531884|2    ....................  ....................  ....................  ....................  ....................  ....................
H    9064 hCoV-19/England/QEUH-960050/2020|EPI_ISL_531885|20    ....................  ....................  ....................  ....................  ....................  ....................
H    9065 hCoV-19/England/QEUH-95FE87/2020|EPI_ISL_531886|20    ....................  ....................  ....................  ....................  ....................  ....................
H    9066 hCoV-19/Scotland/QEUH-9600E7/2020|EPI_ISL_531887|2    ....................  ....................  ....................  ....................  ....................  ....................
H    9067 hCoV-19/England/QEUH-95FC7E/2020|EPI_ISL_531888|20    ....................  ....................  ....................  ....................  ....................  ....................
H    9068 hCoV-19/Scotland/QEUH-95FCF6/2020|EPI_ISL_531889|2    ....................  ....................  ....................  ....................  ....................  ....................
H    9069 hCoV-19/Scotland/QEUH-95FF39/2020|EPI_ISL_531890|2    ....................  ....................  ....................  ....................  ....................  ....................
H    9070 hCoV-19/England/QEUH-95FD02/2020|EPI_ISL_531891|20    ....................  .A..................  ....................  ....................  ....................  ...A................
H    9071 hCoV-19/Scotland/QEUH-95FE78/2020|EPI_ISL_531892|2    ....................  ....................  ....................  ....................  ....................  ....................
H    9072 hCoV-19/England/QEUH-9601E4/2020|EPI_ISL_531893|20    ....................  ....................  ....................  ....................  ....................  ....................
H    9073 hCoV-19/Scotland/QEUH-96023C/2020|EPI_ISL_531894|2    ....................  ....................  ....................  ....................  ....................  ....................
H    9074 hCoV-19/Northern Ireland/QEUH-95FE3C/2020|EPI_ISL_    ....................  ....................  ....................  ....................  ....................  ....................
H    9075 hCoV-19/Northern Ireland/QEUH-95FEF0/2020|EPI_ISL_    ....................  ....................  ....................  ....................  ....................  ....................
H    9076 hCoV-19/England/QEUH-95FD6C/2020|EPI_ISL_531898|20    ....................  ......A.............  ....................  ....................  ....................  ....................
H    9077 hCoV-19/England/QEUH-96020F/2020|EPI_ISL_531899|20    ....................  ....................  ....................  ....................  ....T...............  ....................
H    9078 hCoV-19/South Africa/KRISP-K002479/2020|EPI_ISL_53    ....................  ....................  ....................  ....................  ....................  ....................
H    9079 hCoV-19/South Africa/KRISP-K002484/2020|EPI_ISL_53    ....................  ....................  ....................  ....................  ....................  ....................
H    9080 hCoV-19/South Africa/KRISP-K002486/2020|EPI_ISL_53    ....................  ....................  ....................  ....................  ....................  ....................
H    9081 hCoV-19/South Africa/KRISP-K002490/2020|EPI_ISL_53    ....................  ....................  ....................  ....................  ....................  ....................
H    9082 hCoV-19/South Africa/KRISP-K002491/2020|EPI_ISL_53    ....................  ....................  ....................  ....................  ....................  ....................
H    9083 hCoV-19/South Africa/KRISP-K002495/2020|EPI_ISL_53    ....................  ....................  ....................  ....................  ....................  ....................
H    9084 hCoV-19/South Africa/KRISP-K002498/2020|EPI_ISL_53    ....................  ....................  ....................  ....................  ....................  ....................
H    9085 hCoV-19/South Africa/KRISP-K002501/2020|EPI_ISL_53    ....................  ....................  ....................  ....................  ....................  ....................
H    9086 hCoV-19/South Africa/KRISP-K002502/2020|EPI_ISL_53    ....................  ....................  ....................  ....................  ....................  ....................
```

| | | | | | | | | |
|---|---|---|---|---|---|---|---|---|
| H | 9087 hCoV-19/USA/WI-UW-1231/2020\|EPI_ISL_536604\|2020-08 | ···················· | ···················· | ····················· | ···················· | ···················· | ···················· | |
| H | 9088 hCoV-19/USA/WI-UW-1232/2020\|EPI_ISL_536605\|2020-08 | ···················· | ···················· | ····················· | ···················· | ···················· | ···················· | |
| H | 9089 hCoV-19/USA/WI-UW-1233/2020\|EPI_ISL_536606\|2020-08 | ···················· | ···················· | ····················· | ···················· | ···················· | ···················· | |
| H | 9090 hCoV-19/USA/WI-UW-1234/2020\|EPI_ISL_536607\|2020-08 | ···················· | ···················· | ····················· | ···················· | ···················· | ···················· | |
| H | 9091 hCoV-19/USA/WI-UW-1235/2020\|EPI_ISL_536608\|2020-08 | ···················· | ·A·················· | ····················· | ···················· | ···················· | ···················· | |
| H | 9092 hCoV-19/USA/WI-UW-1236/2020\|EPI_ISL_536609\|2020-08 | ···················· | ···················· | ····················· | ···················· | ···················· | ···················· | |
| H | 9093 hCoV-19/USA/WI-UW-1237/2020\|EPI_ISL_536610\|2020-08 | ···················· | ···················· | ····················· | ···················· | ···················· | ···················· | |
| H | 9094 hCoV-19/USA/WI-UW-1238/2020\|EPI_ISL_536611\|2020-08 | ···················· | ···················· | ····················· | ···················· | ···················· | ···················· | |
| H | 9095 hCoV-19/USA/WI-UW-1239/2020\|EPI_ISL_536612\|2020-08 | ···················· | ···················· | ····················· | ···················· | ···················· | ···················· | |
| H | 9096 hCoV-19/USA/WI-UW-1240/2020\|EPI_ISL_536613\|2020-08 | ···················· | ···················· | ····················· | ···················· | ···················· | ···················· | |
| H | 9097 hCoV-19/USA/WI-UW-1241/2020\|EPI_ISL_536614\|2020-08 | ···················· | ···················· | ····················· | ···················· | ···················· | ···················· | |
| H | 9098 hCoV-19/USA/WI-UW-1242/2020\|EPI_ISL_536615\|2020-08 | ···················· | ···················· | ····················· | ···················· | ···················· | ···················· | |
| H | 9099 hCoV-19/USA/WI-UW-1243/2020\|EPI_ISL_536616\|2020-08 | ···················· | ···················· | ····················· | ···················· | ···················· | ···················· | |
| H | 9100 hCoV-19/USA/WI-UW-1244/2020\|EPI_ISL_536617\|2020-08 | ···················· | ···················· | ····················· | ···················· | ···················· | ···················· | |
| H | 9101 hCoV-19/USA/WI-UW-1245/2020\|EPI_ISL_536618\|2020-08 | ···················· | ···················· | ····················· | ···················· | ···················· | ···················· | |
| H | 9103 hCoV-19/USA/WI-UW-1247/2020\|EPI_ISL_536620\|2020-08 | ···················· | ··················C· | ····················· | ···················· | ···················· | ···················· | 1 |
| H | 9104 hCoV-19/USA/WI-UW-1248/2020\|EPI_ISL_536621\|2020-08 | ···················· | ···················· | ····················· | ···················· | ···················· | ···················· | |
| H | 9105 hCoV-19/USA/WI-UW-1249/2020\|EPI_ISL_536622\|2020-08 | ···················· | ···················· | ····················· | ···················· | ···················· | ···················· | |
| H | 9106 hCoV-19/USA/WI-UW-1251/2020\|EPI_ISL_536624\|2020-08 | ···················· | ···················· | ····················· | ···················· | ···················· | ···················· | |
| H | 9107 hCoV-19/USA/WI-UW-1252/2020\|EPI_ISL_536625\|2020-08 | ···················· | ···················· | ····················· | ···················· | ···················· | ···················· | |
| H | 9108 hCoV-19/USA/FL-BPHL-1268/2020\|EPI_ISL_541200\|2020- | ···················· | ···················· | ····················· | ···················· | ···················· | ···················· | |
| H | 9109 hCoV-19/USA/FL-BPHL-1269/2020\|EPI_ISL_541201\|2020- | ···················· | ···················· | ····················· | ···················· | ···················· | ···················· | |
| H | 9110 hCoV-19/USA/FL-BPHL-1272/2020\|EPI_ISL_541204\|2020- | ···················· | ···················· | ····················· | ···················· | ···················· | ···················· | |
| H | 9111 hCoV-19/USA/FL-BPHL-1273/2020\|EPI_ISL_541205\|2020- | ···················· | ·············A······ | ····················· | ···················· | ···················· | ···················· | |
| H | 9112 hCoV-19/USA/FL-BPHL-1277/2020\|EPI_ISL_541209\|2020- | ···················· | ···················· | ····················· | ···················· | ···················· | ···················· | |
| H | 9113 hCoV-19/USA/FL-BPHL-1278/2020\|EPI_ISL_541210\|2020- | ···················· | ···················· | ····················· | ···················· | ···················· | ···················· | |
| H | 9114 hCoV-19/USA/FL-BPHL-1281/2020\|EPI_ISL_541213\|2020- | ···················· | ···················· | ····················· | ···················· | ···················· | ···················· | |
| H | 9115 hCoV-19/USA/FL-BPHL-1283/2020\|EPI_ISL_541215\|2020- | ···················· | ···················· | ····················· | ···················· | ···················· | ···················· | |
| H | 9116 hCoV-19/USA/FL-BPHL-1285/2020\|EPI_ISL_541217\|2020- | ···················· | ···················· | ····················· | ···················· | ···················· | ···················· | |
| H | 9119 hCoV-19/USA/FL-BPHL-1289/2020\|EPI_ISL_541221\|2020- | ···················· | ···················· | ····················· | ···················· | ···················· | ···················· | |
| H | 9121 hCoV-19/USA/FL-BPHL-1293/2020\|EPI_ISL_541225\|2020- | ···················· | ···················· | ····················· | ···················· | ···················· | ···················· | |
| H | 9122 hCoV-19/USA/FL-BPHL-1294/2020\|EPI_ISL_541226\|2020- | ···················· | ···················· | ····················· | ···················· | ···················· | ···················· | |
| H | 9125 hCoV-19/USA/FL-BPHL-1301/2020\|EPI_ISL_541233\|2020- | ···················· | ···················· | ····················· | ···················· | ···················· | ···················· | |
| H | 9126 hCoV-19/USA/FL-BPHL-1302/2020\|EPI_ISL_541234\|2020- | ···················· | ···················· | ····················· | ···················· | ···················· | ···················· | |
| H | 9128 hCoV-19/USA/FL-BPHL-1227/2020\|EPI_ISL_541237\|2020- | ···················· | ···················· | ····················· | ···················· | ···················· | ···················· | |
| H | 9129 hCoV-19/USA/FL-BPHL-1229/2020\|EPI_ISL_541239\|2020- | ···················· | ···················· | ····················· | ···················· | ···················· | ···················· | |
| H | 9130 hCoV-19/USA/FL-BPHL-1230/2020\|EPI_ISL_541240\|2020- | ···················· | ···················· | ····················· | ···················· | ···················· | ···················· | |
| H | 9131 hCoV-19/USA/FL-BPHL-1231/2020\|EPI_ISL_541241\|2020- | ···················· | ···················· | ····················· | ···················· | ···················· | ···················· | |
| H | 9132 hCoV-19/USA/FL-BPHL-1306/2020\|EPI_ISL_541244\|2020- | ···················· | ···················· | ····················· | ···················· | ···················· | ···················· | |
| H | 9133 hCoV-19/USA/FL-BPHL-1308/2020\|EPI_ISL_541246\|2020- | ···················· | ···················· | ····················· | ···················· | ·NN················· | ···················· | |
| H | 9134 hCoV-19/USA/FL-BPHL-1312/2020\|EPI_ISL_541250\|2020- | ···················· | ···················· | ····················· | ···················· | ···················· | ···················· | |
| H | 9135 hCoV-19/USA/FL-BPHL-1315/2020\|EPI_ISL_541253\|2020- | ···················· | ···················· | ····················· | ···················· | ···················· | ···················· | |
| H | 9136 hCoV-19/USA/FL-BPHL-1317/2020\|EPI_ISL_541255\|2020- | ···················· | ···················· | ····················· | ···················· | ···················· | ···················· | |
| H | 9137 hCoV-19/USA/FL-BPHL-1318/2020\|EPI_ISL_541256\|2020- | ···················· | ···················· | ····················· | ···················· | ···················· | ···················· | |
| H | 9138 hCoV-19/USA/TX-HMH-1026/2020\|EPI_ISL_542500\|2020-0 | ···················· | ···················· | ····················· | ···················· | ···················· | ···················· | |
| H | 9139 hCoV-19/USA/TX-HMH-1035/2020\|EPI_ISL_542502\|2020-0 | ···················· | ···················· | ····················· | ···················· | ···················· | ···················· | |
| H | 9140 hCoV-19/USA/TX-HMH-1036/2020\|EPI_ISL_542503\|2020-0 | ···················· | ···················· | ····················· | ···················· | ···················· | ···················· | |
| H | 9141 hCoV-19/USA/TX-HMH-1041/2020\|EPI_ISL_542504\|2020-0 | ···················· | ···················· | ····················· | ···················· | ···················· | ···················· | |
| H | 9142 hCoV-19/USA/TX-HMH-1044/2020\|EPI_ISL_542505\|2020-0 | ···················· | ···················· | ····················· | ···················· | ···················· | ···················· | |
| H | 9143 hCoV-19/USA/TX-HMH-1045/2020\|EPI_ISL_542506\|2020-0 | ···················· | ···················· | ····················· | ···················· | ···················· | ···················· | |
| H | 9144 hCoV-19/USA/TX-HMH-1046/2020\|EPI_ISL_542507\|2020-0 | ···················· | ···················· | ····················· | ···················· | ···················· | ···················· | |
| H | 9145 hCoV-19/USA/TX-HMH-1052/2020\|EPI_ISL_542508\|2020-0 | ···················· | ···················· | ····················· | ···················· | ···················· | ···················· | |
| H | 9146 hCoV-19/USA/TX-HMH-1054/2020\|EPI_ISL_542509\|2020-0 | ···················· | ···················· | ····················· | ···················· | ···················· | ···················· | |
| H | 9147 hCoV-19/USA/TX-HMH-1056/2020\|EPI_ISL_542510\|2020-0 | ···················· | ···················· | ····················· | ···················· | ···················· | ···················· | |
| H | 9148 hCoV-19/USA/TX-HMH-1057/2020\|EPI_ISL_542511\|2020-0 | ···················· | ···················· | ····················· | ···················· | ···················· | ···················· | |
| H | 9149 hCoV-19/USA/TX-HMH-1058/2020\|EPI_ISL_542512\|2020-0 | ···················· | ···················· | ····················· | ···················· | ···················· | ···················· | |
| H | 9150 hCoV-19/USA/TX-HMH-1059/2020\|EPI_ISL_542513\|2020-0 | ···················· | ···················· | ····················· | ···················· | ···················· | ···················· | |
| H | 9151 hCoV-19/USA/TX-HMH-1060/2020\|EPI_ISL_542514\|2020-0 | ···················· | ···················· | ····················· | ···················· | ···················· | ···················· | |
| H | 9152 hCoV-19/USA/TX-HMH-1066/2020\|EPI_ISL_542515\|2020-0 | ···················· | ···················· | ····················· | ···················· | ···················· | ···················· | |
| H | 9153 hCoV-19/USA/TX-HMH-1069/2020\|EPI_ISL_542516\|2020-0 | ···················· | ···················· | ····················· | ···················· | ···················· | ···················· | |
| H | 9154 hCoV-19/USA/TX-HMH-1070/2020\|EPI_ISL_542517\|2020-0 | ···················· | ···················· | ····················· | ···················· | ···················· | ···················· | |
| H | 9155 hCoV-19/USA/TX-HMH-1071/2020\|EPI_ISL_542518\|2020-0 | ···················· | ···················· | ····················· | ···················· | ···················· | ···················· | |
| H | 9156 hCoV-19/USA/TX-HMH-1072/2020\|EPI_ISL_542519\|2020-0 | ···················· | ···················· | ····················· | ···················· | ···················· | ···················· | |
| H | 9157 hCoV-19/USA/TX-HMH-1073/2020\|EPI_ISL_542520\|2020-0 | ···················· | ···················· | ····················· | ···················· | ···················· | ···················· | |
| H | 9158 hCoV-19/USA/TX-HMH-1074/2020\|EPI_ISL_542521\|2020-0 | ···················· | ···················· | ····················· | ···················· | ···················· | ···················· | |
| H | 9159 hCoV-19/USA/TX-HMH-1075/2020\|EPI_ISL_542522\|2020-0 | ···················· | ···················· | ····················· | ···················· | ···················· | ···················· | |
| H | 9161 hCoV-19/USA/TX-HMH-1083/2020\|EPI_ISL_542524\|2020-0 | ···················· | ···················· | ····················· | ···················· | ···················· | ···················· | |
| H | 9162 hCoV-19/USA/TX-HMH-1089/2020\|EPI_ISL_542526\|2020-0 | ···················· | ···················· | ····················· | ···················· | ···················· | ···················· | |
| H | 9163 hCoV-19/USA/TX-HMH-1092/2020\|EPI_ISL_542527\|2020-0 | ···················· | ···················· | ····················· | ···················· | ···················· | ···················· | |
| H | 9164 hCoV-19/USA/TX-HMH-1093/2020\|EPI_ISL_542528\|2020-0 | ···················· | ··············C····· | ····················· | ···················· | ···················· | ···················· | |
| H | 9166 hCoV-19/USA/TX-HMH-1100/2020\|EPI_ISL_542530\|2020-0 | ···················· | ···················· | ····················· | ···················· | ···················· | ···················· | |
| H | 9168 hCoV-19/USA/TX-HMH-1103/2020\|EPI_ISL_542532\|2020-0 | ···················· | ···················· | ····················· | ···················· | ···················· | ···················· | |
| H | 9169 hCoV-19/USA/TX-HMH-1104/2020\|EPI_ISL_542533\|2020-0 | ···················· | ···················· | ····················· | ···················· | ···················· | ···················· | |
| H | 9170 hCoV-19/USA/TX-HMH-1108/2020\|EPI_ISL_542534\|2020-0 | ···················· | ···················· | ····················· | ···················· | ···················· | ···················· | |
| H | 9171 hCoV-19/USA/TX-HMH-1109/2020\|EPI_ISL_542535\|2020-0 | ···················· | ···················· | ····················· | ···················· | ···················· | ···················· | |
| H | 9172 hCoV-19/USA/TX-HMH-1110/2020\|EPI_ISL_542536\|2020-0 | ···················· | ···················· | ····················· | ···················· | ···················· | ···················· | |
| H | 9173 hCoV-19/USA/TX-HMH-1111/2020\|EPI_ISL_542537\|2020-0 | ···················· | ···················· | ····················· | ···················· | ···················· | ···················· | |
| H | 9175 hCoV-19/USA/TX-HMH-1113/2020\|EPI_ISL_542539\|2020-0 | ···················· | ···················· | ····················· | ···················· | ···················· | ···A················ | |
| H | 9178 hCoV-19/USA/TX-HMH-1120/2020\|EPI_ISL_542542\|2020-0 | ···················· | ···················· | ····················· | ···················· | ···················· | ···················· | |
| H | 9179 hCoV-19/USA/TX-HMH-1125/2020\|EPI_ISL_542543\|2020-0 | ···················· | ···················· | ····················· | ···················· | ···················· | ···················· | |

```
H 9180 hCoV-19/USA/TX-HMH-1127/2020|EPI_ISL_542544|2020-0 ···················· ···················· ····················· ···················· ···················· ····················
H 9182 hCoV-19/USA/TX-HMH-1137/2020|EPI_ISL_542546|2020-0 ···················· ···················· ····················· ···················· ···················· ····················
H 9183 hCoV-19/USA/TX-HMH-1139/2020|EPI_ISL_542547|2020-0 ···················· ···················· ····················· ···················· ···················· ····················
H 9185 hCoV-19/USA/TX-HMH-1142/2020|EPI_ISL_542549|2020-0 ···················· ···················· ····················· ···················· ···················· ····················
H 9187 hCoV-19/USA/TX-HMH-1144/2020|EPI_ISL_542551|2020-0 ···················· ···················· ····················· ···················· ···················· ····················
H 9189 hCoV-19/USA/TX-HMH-1148/2020|EPI_ISL_542553|2020-0 ···················· ···················· ····················· ···················· ···················· ····················
H 9190 hCoV-19/USA/TX-HMH-1155/2020|EPI_ISL_542556|2020-0 ···················· ···················· ····················· ···················· ···················· ····················
H 9191 hCoV-19/USA/TX-HMH-1156/2020|EPI_ISL_542557|2020-0 ···················· ···················· ····················· ···················· ···················· ····················
H 9194 hCoV-19/USA/TX-HMH-1162/2020|EPI_ISL_542561|2020-0 ···················· ···················· ····················· ···················· ···················· ····················
H 9195 hCoV-19/USA/TX-HMH-1164/2020|EPI_ISL_542562|2020-0 ···················· ···················· ····················· ···················· ···················· ····················
H 9196 hCoV-19/USA/TX-HMH-1175/2020|EPI_ISL_542565|2020-0 ···················· ···················· ····················· ···················· ···················· ····················
H 9198 hCoV-19/USA/TX-HMH-1179/2020|EPI_ISL_542568|2020-0 ···················· ···················· ····················· ···················· ···················· ····················
H 9199 hCoV-19/USA/TX-HMH-1182/2020|EPI_ISL_542570|2020-0 ···················· ···················· ····················· ···················· ···················· ····················
H 9200 hCoV-19/USA/TX-HMH-1183/2020|EPI_ISL_542571|2020-0 ···················· ···················· ····················· ···················· ···················· ····················
H 9201 hCoV-19/USA/TX-HMH-1184/2020|EPI_ISL_542572|2020-0 ···················· ···················· ····················· ···················· ···················· ····················
H 9202 hCoV-19/USA/TX-HMH-1185/2020|EPI_ISL_542573|2020-0 ···················· ···················· ····················· ···················· ···················· ····················
H 9203 hCoV-19/USA/TX-HMH-1186/2020|EPI_ISL_542574|2020-0 ···················· ···················· ····················· ···················· ···················· ····················
H 9204 hCoV-19/USA/TX-HMH-1187/2020|EPI_ISL_542575|2020-0 ···················· ···················· ····················· ···················· ···················· ····················
H 9205 hCoV-19/USA/TX-HMH-1188/2020|EPI_ISL_542576|2020-0 ···················· ···················· ····················· ···················· ···················· ····················
H 9206 hCoV-19/USA/TX-HMH-1189/2020|EPI_ISL_542577|2020-0 ···················· ···················· ····················· ···················· ···················· ····················
H 9207 hCoV-19/USA/TX-HMH-1190/2020|EPI_ISL_542578|2020-0 ···················· ···················· ····················· ···················· ···················· ····················
H 9208 hCoV-19/USA/TX-HMH-1194/2020|EPI_ISL_542579|2020-0 ···················· ···················· ····················· ···················· ···················· ····················
H 9209 hCoV-19/USA/TX-HMH-1195/2020|EPI_ISL_542580|2020-0 ···················· ···················· ····················· ···················· ···················· ····················
H 9210 hCoV-19/USA/TX-HMH-1197/2020|EPI_ISL_542581|2020-0 ···················· ···················· ····················· ···················· ···················· ····················
H 9211 hCoV-19/USA/TX-HMH-1199/2020|EPI_ISL_542582|2020-0 ···················· ···················· ····················· ···················· ···················· ····················
H 9212 hCoV-19/USA/TX-HMH-1200/2020|EPI_ISL_542583|2020-0 ···················· ···················· ····················· ···················· ···················· ····················
H 9213 hCoV-19/USA/TX-HMH-1201/2020|EPI_ISL_542584|2020-0 ···················· ···················· ····················· ···················· ···················· ····················
H 9214 hCoV-19/USA/TX-HMH-1205/2020|EPI_ISL_542585|2020-0 ···················· ···················· ····················· ···················· ···················· ····················
H 9215 hCoV-19/USA/TX-HMH-1210/2020|EPI_ISL_542586|2020-0 ···················· ···················· ····················· ···················· ···················· ····················
H 9216 hCoV-19/USA/TX-HMH-1213/2020|EPI_ISL_542587|2020-0 ···················· ···················· ····················· ···················· ···················· ····················
H 9217 hCoV-19/England/QEUH-96EE85/2020|EPI_ISL_530501|20 ···················· ···················· ····················· ···················· ···················· ····················
H 9218 hCoV-19/England/QEUH-96E9A2/2020|EPI_ISL_530503|20 ···················· ···················· ····················· ···················· ···················· ····················
H 9219 hCoV-19/England/QEUH-96F428/2020|EPI_ISL_530504|20 ···················· ···················· ····················· ···················· ···················· ····················
H 9220 hCoV-19/England/QEUH-96F1C7/2020|EPI_ISL_530505|20 ···················· ···················· ····················· ···················· ···················· ····················
H 9221 hCoV-19/Northern Ireland/QEUH-96EC7C/2020|EPI_ISL_ ···················· ···················· ····················· ···················· ···················· ····················
H 9222 hCoV-19/England/QEUH-96E623/2020|EPI_ISL_530507|20 ···················· ···················· ····················· ···················· ···················· ····················
H 9223 hCoV-19/England/QEUH-96E474/2020|EPI_ISL_530508|20 ···················· ···················· ····················· ···················· ···················· ····················
H 9224 hCoV-19/England/QEUH-96DF74/2020|EPI_ISL_530511|20 ···················· ···················· ····················· ···················· ···················· ····················
H 9225 hCoV-19/Northern Ireland/QEUH-96ECF4/2020|EPI_ISL_ ···················· ···················· ····················· ···················· ···················· ····················
H 9226 hCoV-19/England/QEUH-96F15E/2020|EPI_ISL_530514|20 ···················· ···················· ····················· ···················· ···················· ····················
H 9227 hCoV-19/England/QEUH-96EBAC/2020|EPI_ISL_530515|20 ···················· ···················· ····················· ···················· ···················· ····················
H 9228 hCoV-19/England/QEUH-96F2B5/2020|EPI_ISL_530516|20 ···················· ···················· ····················· ···················· ···················· ····················
H 9229 hCoV-19/Northern Ireland/QEUH-96EDF1/2020|EPI_ISL_ ···················· ···················· ····················· ···················· ···················· ····················
H 9230 hCoV-19/England/QEUH-96DED1/2020|EPI_ISL_530518|20 ···················· ···················· ····················· ···················· ···················· ····················
H 9231 hCoV-19/England/QEUH-96E650/2020|EPI_ISL_530519|20 ···················· ···················· ····················· ···················· ······T············· ····················
H 9232 hCoV-19/England/QEUH-96EE67/2020|EPI_ISL_530520|20 ···················· ···················· ····················· ···················· ···················· ····················
H 9233 hCoV-19/Northern Ireland/QEUH-96215B/2020|EPI_ISL_ ···················· ···················· ····················· ···················· ···················· ····················
H 9234 hCoV-19/Northern Ireland/QEUH-962188/2020|EPI_ISL_ ···················· ···················· ····················· ···················· ···················· ····················
H 9235 hCoV-19/England/QEUH-962D1B/2020|EPI_ISL_531810|20 ···················· ···················· ····················· ···················· ···················· ····················
H 9236 hCoV-19/England/QEUH-963846/2020|EPI_ISL_531811|20 ···················· ···················· ····················· ···················· ···················· ····················
H 9237 hCoV-19/Scotland/QEUH-962CF0/2020|EPI_ISL_531814|2 ···················· ···················· ····················· ···················· ···················· ····················
H 9238 hCoV-19/Scotland/QEUH-962C0F/2020|EPI_ISL_531815|2 ···················· ···················· ····················· ···················· ···················· ····················
H 9239 hCoV-19/England/QEUH-9633EB/2020|EPI_ISL_531816|20 ···················· ···················· ····················· ···················· ···················· ····················
H 9240 hCoV-19/England/QEUH-963794/2020|EPI_ISL_531818|20 ···················· ···················· ····················· ···················· ···················· ····················
H 9241 hCoV-19/England/QEUH-962E90/2020|EPI_ISL_531819|20 ···················· ·A·················· ····················· ···················· ···················· ··A·················
H 9242 hCoV-19/England/QEUH-962689/2020|EPI_ISL_531820|20 ···················· ···················· ····················· ···················· ···················· ····················
H 9243 hCoV-19/Scotland/QEUH-962EDC/2020|EPI_ISL_531821|2 ···················· ···················· ····················· ···················· ···················· ····················
H 9244 hCoV-19/England/QEUH-963381/2020|EPI_ISL_531822|20 ···················· ···················· ····················· ···················· ···················· ····················
H 9245 hCoV-19/Scotland/QEUH-962328/2020|EPI_ISL_531824|2 ···················· ···················· ····················· ···················· ···················· ····················
H 9246 hCoV-19/England/QEUH-962601/2020|EPI_ISL_531825|20 ···················· ···················· ····················· ···················· ···················· ····················
H 9247 hCoV-19/Northern Ireland/QEUH-9621B5/2020|EPI_ISL_ ···················· ···················· ····················· ···················· ···················· ····················
H 9248 hCoV-19/Northern Ireland/QEUH-963284/2020|EPI_ISL_ ···················· ···················· ····················· ···················· ···················· ····················
H 9249 hCoV-19/Wales/QEUH-961F8F/2020|EPI_ISL_531830|2020 ···················· ···················· ····················· ···················· ···················· ····················
H 9250 hCoV-19/England/QEUH-96356D/2020|EPI_ISL_531831|20 ···················· ···················· ····················· ···················· ···················· ····················
H 9251 hCoV-19/Scotland/QEUH-963345/2020|EPI_ISL_531832|2 ···················· ···················· ····················· ···················· ···················· ····················
H 9252 hCoV-19/England/QEUH-96263E/2020|EPI_ISL_531833|20 ···················· ···················· ····················· ···················· ···················· ····················
H 9253 hCoV-19/Northern Ireland/QEUH-963099/2020|EPI_ISL_ ···················· ···················· ····················· ···················· ···················· ····················
H 9254 hCoV-19/England/QEUH-96248F/2020|EPI_ISL_531836|20 ···················· ···················· ····················· ···················· ···················· ····················
H 9255 hCoV-19/England/QEUH-962EAF/2020|EPI_ISL_531837|20 ···················· ·A·················· ····················· ···················· ···················· ··A·················
H 9256 hCoV-19/England/QEUH-962CB4/2020|EPI_ISL_531838|20 ···················· ···················· ····················· ···················· ···················· ····················
H 9257 hCoV-19/England/QEUH-962F42/2020|EPI_ISL_531839|20 ···················· ···················· ····················· ···················· ···················· ····················
H 9258 hCoV-19/Northern Ireland/QEUH-96204F/2020|EPI_ISL_ ···················· ···················· ····················· ···················· ···················· ····················
H 9259 hCoV-19/England/QEUH-962504/2020|EPI_ISL_531841|20 ···················· ···················· ····················· ···················· ···················· ····················
H 9260 hCoV-19/Scotland/QEUH-962B20/2020|EPI_ISL_531842|2 ···················· ···················· ····················· ···················· ···················· ····················
H 9261 hCoV-19/Northern Ireland/QEUH-962285/2020|EPI_ISL_ ···················· ···················· ····················· ···················· ···················· ····················
H 9262 hCoV-19/England/QEUH-9635E5/2020|EPI_ISL_531844|20 ···················· ·A······CA·········· ····················· ···················· ···················· ···················· 1
H 9263 hCoV-19/Northern Ireland/QEUH-9620E5/2020|EPI_ISL_ ···················· ···················· ····················· ···················· ···················· ····················
H 9264 hCoV-19/England/QEUH-9623DD/2020|EPI_ISL_531846|20 ···················· ···················· ····················· ···················· ···················· ····················
H 9265 hCoV-19/Northern Ireland/QEUH-962443/2020|EPI_ISL_ ···················· ···················· ····················· ···················· ···················· ····················
```

H 9266 hCoV-19/England/QEUH-962407/2020|EPI_ISL_531849|20 .................... .................... .................... .................... .................... ....................

Group 3 - Sep 2020-Oct 2020

| A | B | C | D | E | F | G | H | I | J | K | L |
|---|---|---|---|---|---|---|---|---|---|---|---|
| Database | Database ID | GISAID ID | GCTGCAATCGTGCTACAACT | TGAACTGTTGCGACTACGTG | TTCGGAAGAGACAGGTACGTT | CACACAATCGAAGCGCAGTA | GCTGGTGCTGCAGCTTATTA | AGGGTCAAGTGCACAGTCTA | mismatches? | mismatches? | primer set? |
| I | 2 | hCoV-19/USA/TX-HMH-3319/2020\|EPI_ISL_544129\|2020-0 | .................... | .................... | .................... | .................... | .................... | .................... | | | |
| I | 3 | hCoV-19/USA/TX-HMH-3321/2020\|EPI_ISL_544130\|2020-0 | .................... | .................... | .................... | .................... | .................... | .................... | | | |
| I | 4 | hCoV-19/USA/TX-HMH-3322/2020\|EPI_ISL_544131\|2020-0 | .................... | .................... | .................... | .................... | .................... | .................... | | | |
| I | 5 | hCoV-19/USA/TX-HMH-3324/2020\|EPI_ISL_544133\|2020-0 | .................... | .................... | .................... | .................... | .................... | .................... | | | |
| I | 6 | hCoV-19/USA/TX-HMH-3325/2020\|EPI_ISL_544134\|2020-0 | .................... | .................... | .................... | .................... | .................... | .................... | | | |
| I | 7 | hCoV-19/USA/TX-HMH-3352/2020\|EPI_ISL_544137\|2020-0 | .................... | .................... | .................... | .................... | .................... | .................... | | | |
| I | 8 | hCoV-19/USA/TX-HMH-3353/2020\|EPI_ISL_544138\|2020-0 | .................... | .................... | .................... | .................... | .................... | .................... | | | |
| I | 9 | hCoV-19/USA/TX-HMH-3355/2020\|EPI_ISL_544140\|2020-0 | .................... | .................... | .................... | .................... | .................... | .................... | | | |
| I | 10 | hCoV-19/USA/TX-HMH-3357/2020\|EPI_ISL_544142\|2020-0 | .................... | .................... | .................... | .................... | .................... | .................... | | | |
| I | 12 | hCoV-19/USA/TX-HMH-3359/2020\|EPI_ISL_544144\|2020-0 | .................... | .................... | .................... | .................... | .................... | .................... | | | |
| I | 13 | hCoV-19/USA/TX-HMH-3360/2020\|EPI_ISL_544145\|2020-0 | .................... | .................... | .................... | .................... | .................... | .................... | | | |
| I | 14 | hCoV-19/USA/TX-HMH-3361/2020\|EPI_ISL_544146\|2020-0 | .................... | .................... | .................... | .................... | .................... | .................... | | | |
| I | 15 | hCoV-19/USA/TX-HMH-3362/2020\|EPI_ISL_544147\|2020-0 | .................... | .................... | .................... | .................... | .................... | .................... | | | |
| I | 16 | hCoV-19/USA/TX-HMH-3363/2020\|EPI_ISL_544148\|2020-0 | .................... | .................... | .................... | .................... | .................... | .................... | | | |
| I | 17 | hCoV-19/USA/TX-HMH-3366/2020\|EPI_ISL_544150\|2020-0 | .................... | .................... | .................... | .................... | .................... | .................... | | | |
| I | 18 | hCoV-19/USA/TX-HMH-3367/2020\|EPI_ISL_544151\|2020-0 | .................... | .................... | .................... | .................... | ..............N..... | .................... | | | |
| I | 19 | hCoV-19/USA/TX-HMH-3369/2020\|EPI_ISL_544153\|2020-0 | .................... | .................... | .................... | .................... | .................... | .................... | | | |
| I | 20 | hCoV-19/USA/TX-HMH-3370/2020\|EPI_ISL_544154\|2020-0 | .................... | .................... | .................... | .................... | .................... | .................... | | | |
| I | 21 | hCoV-19/USA/TX-HMH-3371/2020\|EPI_ISL_544155\|2020-0 | .................... | .................... | .................... | .................... | .................... | .................... | | | |
| I | 22 | hCoV-19/USA/TX-HMH-3372/2020\|EPI_ISL_544156\|2020-0 | .................... | .................... | .................... | .................... | .................... | .................... | | | |
| I | 23 | hCoV-19/USA/TX-HMH-3374/2020\|EPI_ISL_544158\|2020-0 | .................... | .................... | .................... | .................... | .................... | .................... | | | |
| I | 25 | hCoV-19/USA/TX-HMH-3400/2020\|EPI_ISL_544160\|2020-0 | .................... | .................... | .................... | .................... | .................... | .................... | | | |
| I | 26 | hCoV-19/USA/TX-HMH-3401/2020\|EPI_ISL_544161\|2020-0 | .................... | .................... | .................... | .................... | .................... | .................... | | | |
| I | 27 | hCoV-19/USA/TX-HMH-3402/2020\|EPI_ISL_544162\|2020-0 | .................... | .................... | .................... | .................... | .................... | .................... | | | |
| I | 29 | hCoV-19/USA/TX-HMH-3404/2020\|EPI_ISL_544164\|2020-0 | .................... | .................... | .................... | .................... | .................... | .................... | | | |
| I | 30 | hCoV-19/USA/TX-HMH-3405/2020\|EPI_ISL_544165\|2020-0 | .................... | .................... | .................... | .................... | .................... | .................... | | | |
| I | 31 | hCoV-19/USA/TX-HMH-3406/2020\|EPI_ISL_544166\|2020-0 | .................... | .................... | .................... | .................... | .................... | .................... | | | |
| I | 32 | hCoV-19/USA/TX-HMH-3407/2020\|EPI_ISL_544167\|2020-0 | .................... | .................... | .................... | .................... | .................... | .................... | | | |
| I | 36 | hCoV-19/USA/TX-HMH-3412/2020\|EPI_ISL_544172\|2020-0 | .................... | .............A...... | .................... | .................... | .................... | .................... | | | |
| I | 39 | hCoV-19/USA/TX-HMH-3417/2020\|EPI_ISL_544176\|2020-0 | .................... | .................... | .................... | .................... | .................... | .................... | | | |
| I | 40 | hCoV-19/USA/TX-HMH-3420/2020\|EPI_ISL_544179\|2020-0 | .................... | .................... | .................... | .................... | .................... | .................... | | | |
| I | 42 | hCoV-19/USA/TX-HMH-3424/2020\|EPI_ISL_544182\|2020-0 | .................... | .................... | .................... | .................... | .................... | .................... | | | |
| I | 43 | hCoV-19/USA/TX-HMH-3427/2020\|EPI_ISL_544184\|2020-0 | .................... | .................... | .................... | .................... | .................... | .................... | | | |
| I | 44 | hCoV-19/USA/TX-HMH-3428/2020\|EPI_ISL_544185\|2020-0 | .................... | .................... | .................... | .................... | .................... | .................... | | | |
| I | 45 | hCoV-19/USA/TX-HMH-3429/2020\|EPI_ISL_544186\|2020-0 | .................... | .............A...... | .................... | .................... | .................... | .................... | | | |
| I | 46 | hCoV-19/USA/TX-HMH-3431/2020\|EPI_ISL_544188\|2020-0 | .................... | .................... | .................... | .................... | .................... | .................... | | | |
| I | 47 | hCoV-19/USA/TX-HMH-3433/2020\|EPI_ISL_544190\|2020-0 | .................... | .................... | .................... | .................... | .................... | .................... | | | |
| I | 48 | hCoV-19/USA/TX-HMH-3434/2020\|EPI_ISL_544191\|2020-0 | .................... | .................... | .................... | .................... | .................... | N................... | | | |
| I | 49 | hCoV-19/USA/TX-HMH-3436/2020\|EPI_ISL_544192\|2020-0 | .................... | .............A...... | .................... | .................... | .................... | .................... | | | |
| I | 50 | hCoV-19/USA/TX-HMH-3438/2020\|EPI_ISL_544194\|2020-0 | .................... | .................... | .................... | .................... | .................... | .................... | | | |
| I | 52 | hCoV-19/USA/TX-HMH-3441/2020\|EPI_ISL_544196\|2020-0 | .................... | .................... | .................... | .................... | .................... | .................... | | | |
| I | 53 | hCoV-19/USA/TX-HMH-3442/2020\|EPI_ISL_544197\|2020-0 | .................... | .................... | .................... | .................... | .................... | .................... | | | |
| I | 55 | hCoV-19/USA/TX-HMH-2577/2020\|EPI_ISL_545400\|2020-0 | .................... | .................... | .................... | .................... | .................... | .................... | | | |
| I | 56 | hCoV-19/USA/TX-HMH-2578/2020\|EPI_ISL_545401\|2020-0 | .................... | .................... | .................... | .................... | .................... | .................... | | | |
| I | 57 | hCoV-19/USA/TX-HMH-2579/2020\|EPI_ISL_545402\|2020-0 | .................... | ......A............. | .................... | .................... | .................... | .................... | | | |
| I | 58 | hCoV-19/USA/TX-HMH-2582/2020\|EPI_ISL_545405\|2020-0 | .................... | .................... | .................... | .................... | ...............N.... | .................... | | | |
| I | 59 | hCoV-19/USA/TX-HMH-2583/2020\|EPI_ISL_545406\|2020-0 | .................... | .................... | .................... | .................... | .................... | .................... | | | |
| I | 60 | hCoV-19/USA/TX-HMH-2584/2020\|EPI_ISL_545407\|2020-0 | .................... | ......A............. | .................... | .................... | .................... | .................... | | | |
| I | 61 | hCoV-19/USA/TX-HMH-2586/2020\|EPI_ISL_545409\|2020-0 | .................... | .................... | .................... | .................... | .................... | .................... | | | |
| I | 62 | hCoV-19/USA/TX-HMH-2588/2020\|EPI_ISL_545411\|2020-0 | .................... | .................... | .................... | .................... | .................... | .................... | | | |
| I | 64 | hCoV-19/USA/TX-HMH-2590/2020\|EPI_ISL_545413\|2020-0 | .................... | .................... | .................... | .................... | .................... | .................... | | | |
| I | 65 | hCoV-19/USA/TX-HMH-2591/2020\|EPI_ISL_545414\|2020-0 | .................... | .................... | .................... | .................... | ..............N..... | .................... | | | |
| I | 66 | hCoV-19/USA/TX-HMH-2595/2020\|EPI_ISL_545416\|2020-0 | .................... | .................... | .................... | .................... | .................... | .................... | | | |
| I | 67 | hCoV-19/USA/TX-HMH-2602/2020\|EPI_ISL_545419\|2020-0 | .................... | .................... | .................... | .................... | .................... | .................... | | | |
| I | 68 | hCoV-19/USA/TX-HMH-2603/2020\|EPI_ISL_545420\|2020-0 | .................... | .................... | .................... | .................... | .................... | .................... | | | |
| I | 69 | hCoV-19/USA/TX-HMH-2605/2020\|EPI_ISL_545421\|2020-0 | .................... | .................... | .................... | .................... | .................... | .................... | | | |
| I | 70 | hCoV-19/USA/TX-HMH-2606/2020\|EPI_ISL_545422\|2020-0 | .................... | .................... | .................... | .................... | .................... | .................... | | | |
| I | 71 | hCoV-19/USA/TX-HMH-2607/2020\|EPI_ISL_545423\|2020-0 | .................... | .................... | .................... | .................... | .................... | .................... | | | |
| I | 72 | hCoV-19/USA/TX-HMH-2608/2020\|EPI_ISL_545424\|2020-0 | .................... | .................... | .................... | .................... | .................... | .................... | | | |
| I | 73 | hCoV-19/USA/TX-HMH-2609/2020\|EPI_ISL_545425\|2020-0 | .................... | .................... | .................... | .................... | .................... | .................... | | | |
| I | 74 | hCoV-19/USA/TX-HMH-2617/2020\|EPI_ISL_545430\|2020-0 | .................... | .................... | .................... | .................... | .................... | .................... | | | |
| I | 75 | hCoV-19/USA/TX-HMH-2664/2020\|EPI_ISL_545461\|2020-0 | .................... | .................... | .................... | .................... | .................... | .................... | | | |
| I | 76 | hCoV-19/USA/TX-HMH-2666/2020\|EPI_ISL_545462\|2020-0 | .................... | .............A...... | .................... | .................... | .................... | .................... | | | |
| I | 77 | hCoV-19/USA/TX-HMH-2667/2020\|EPI_ISL_545463\|2020-0 | .................... | ......A............. | .................... | .................... | .................... | .................... | | | |
| I | 78 | hCoV-19/USA/TX-HMH-2668/2020\|EPI_ISL_545464\|2020-0 | .................... | .................... | .................... | .................... | .................... | .................... | | | |
| I | 79 | hCoV-19/USA/TX-HMH-2672/2020\|EPI_ISL_545466\|2020-0 | .................... | .................... | .................... | .................... | .................... | .................... | | | |
| I | 81 | hCoV-19/USA/TX-HMH-2674/2020\|EPI_ISL_545468\|2020-0 | .................... | .................... | .................... | .................... | .................... | .................... | | | |
| I | 82 | hCoV-19/USA/TX-HMH-2675/2020\|EPI_ISL_545469\|2020-0 | .................... | .................... | .................... | .................... | .................... | .................... | | | |
| I | 83 | hCoV-19/USA/TX-HMH-2676/2020\|EPI_ISL_545470\|2020-0 | .................... | .................... | .................... | .................... | .................... | .................... | | | |

| | | | | | | | | | |
|---|---|---|---|---|---|---|---|---|---|
| I | 84 | hCoV-19/USA/TX-HMH-2679/2020\|EPI_ISL_545471\|2020-0 | ···················· | ···················· | ····················· | ···················· | ···················· | NN·················· | |
| I | 85 | hCoV-19/USA/TX-HMH-2682/2020\|EPI_ISL_545472\|2020-0 | ···················· | ···················· | ····················· | ···················· | ···················· | ···················· | |
| I | 86 | hCoV-19/USA/TX-HMH-2686/2020\|EPI_ISL_545475\|2020-0 | ···················· | ···················· | ····················· | ···················· | ···················· | ···················· | |
| I | 87 | hCoV-19/USA/TX-HMH-2687/2020\|EPI_ISL_545476\|2020-0 | ···················· | ···················· | ····················· | ···················· | ···················· | ···················· | |
| I | 88 | hCoV-19/USA/TX-HMH-2688/2020\|EPI_ISL_545477\|2020-0 | ···················· | ···················· | ····················· | ···················· | ···················· | ···················· | |
| I | 89 | hCoV-19/USA/TX-HMH-2689/2020\|EPI_ISL_545478\|2020-0 | ···················· | ···················· | ····················· | ···················· | ···················· | ···················· | |
| I | 90 | hCoV-19/USA/TX-HMH-2691/2020\|EPI_ISL_545479\|2020-0 | ···················· | ···················· | ····················· | ···················· | ···················· | ···················· | |
| I | 91 | hCoV-19/USA/TX-HMH-2692/2020\|EPI_ISL_545480\|2020-0 | ···················· | ··············C····· | ····················· | ···················· | ···················· | ···················· | |
| I | 92 | hCoV-19/USA/TX-HMH-2694/2020\|EPI_ISL_545482\|2020-0 | ···················· | ···················· | ····················· | ···················· | ···················· | ···················· | |
| I | 93 | hCoV-19/USA/TX-HMH-2695/2020\|EPI_ISL_545483\|2020-0 | ···················· | ···················· | ····················· | ···················· | ···················· | ···················· | |
| I | 94 | hCoV-19/USA/TX-HMH-2697/2020\|EPI_ISL_545484\|2020-0 | ···················· | ···················· | ····················· | ···················· | ···················· | ···················· | |
| I | 95 | hCoV-19/USA/TX-HMH-2698/2020\|EPI_ISL_545485\|2020-0 | ···················· | ···················· | ····················· | ···················· | ···················· | ···················· | |
| I | 96 | hCoV-19/USA/TX-HMH-2700/2020\|EPI_ISL_545487\|2020-0 | ···················· | ···················· | ····················· | ···················· | ···················· | ···················· | |
| I | 97 | hCoV-19/USA/TX-HMH-2703/2020\|EPI_ISL_545489\|2020-0 | ···················· | ···················· | ····················· | ···················· | ···················· | ···················· | |
| I | 98 | hCoV-19/USA/TX-HMH-2704/2020\|EPI_ISL_545490\|2020-0 | ···················· | ··············C····· | ····················· | ···················· | ···················· | ···················· | |
| I | 99 | hCoV-19/USA/TX-HMH-2705/2020\|EPI_ISL_545491\|2020-0 | ···················· | ···················· | ····················· | ···················· | ···················· | ···················· | |
| I | 100 | hCoV-19/USA/TX-HMH-2706/2020\|EPI_ISL_545492\|2020-0 | ···················· | ···················· | ····················· | ···················· | ···················· | ···················· | |
| I | 103 | hCoV-19/USA/TX-HMH-2713/2020\|EPI_ISL_545497\|2020-0 | ···················· | ·············A······ | ····················· | ···················· | ···················· | ···················· | |
| I | 104 | hCoV-19/USA/TX-HMH-2715/2020\|EPI_ISL_545498\|2020-0 | ···················· | ···················· | ····················· | ···················· | ···················· | ···················· | |
| I | 105 | hCoV-19/USA/TX-HMH-7357/2020\|EPI_ISL_546702\|2020-0 | ···················· | ···················· | ····················· | ···················· | ···················· | ···················· | |
| I | 106 | hCoV-19/USA/TX-HMH-7359/2020\|EPI_ISL_546704\|2020-0 | ···················· | ···················· | ····················· | ···················· | ···················· | ···················· | |
| I | 107 | hCoV-19/USA/TX-HMH-7360/2020\|EPI_ISL_546706\|2020-0 | ···················· | ···················· | ····················· | ···················· | ···················· | ···················· | |
| I | 108 | hCoV-19/USA/TX-HMH-7361/2020\|EPI_ISL_546707\|2020-0 | ···················· | ···················· | ····················· | ···················· | ···················· | ···················· | |
| I | 109 | hCoV-19/USA/TX-HMH-7364/2020\|EPI_ISL_546709\|2020-0 | ···················· | ···················· | ····················· | ···················· | ···················· | ··A················· | |
| I | 110 | hCoV-19/USA/TX-HMH-7366/2020\|EPI_ISL_546710\|2020-0 | ···················· | ·A·················· | ····················· | ···················· | ···················· | ···················· | |
| I | 111 | hCoV-19/USA/TX-HMH-7368/2020\|EPI_ISL_546711\|2020-0 | ···················· | ·············A······ | ····················· | ···················· | ···················· | ···················· | |
| I | 112 | hCoV-19/USA/TX-HMH-7369/2020\|EPI_ISL_546712\|2020-0 | ···················· | ···················· | ····················· | ···················· | ···················· | ···················· | |
| I | 113 | hCoV-19/USA/TX-HMH-7371/2020\|EPI_ISL_546713\|2020-0 | ···················· | ···················· | ····················· | ···················· | ···················· | ···················· | |
| I | 114 | hCoV-19/USA/TX-HMH-7372/2020\|EPI_ISL_546714\|2020-0 | ···················· | ···················· | ····················· | ···················· | ···················· | ···················· | |
| I | 117 | hCoV-19/USA/TX-HMH-7379/2020\|EPI_ISL_546718\|2020-0 | ···················· | ··············C····· | ····················· | ···················· | ··············N····· | N··················· | |
| I | 118 | hCoV-19/USA/TX-HMH-738/2020\|EPI_ISL_546719\|2020-04 | ···················· | ···················· | ····················· | ···················· | ···················· | ···················· | |
| I | 119 | hCoV-19/USA/TX-HMH-7382/2020\|EPI_ISL_546721\|2020-0 | ···················· | ·············A······ | ····················· | ···················· | ··············NN···· | ·······NN··········· | |
| I | 120 | hCoV-19/USA/TX-HMH-7383/2020\|EPI_ISL_546722\|2020-0 | ···················· | ·A·················· | ····················· | ···················· | ···················· | ···················· | |
| I | 121 | hCoV-19/USA/TX-HMH-7384/2020\|EPI_ISL_546723\|2020-0 | ···················· | ···················· | ····················· | ···················· | ···················· | ···················· | |
| I | 125 | hCoV-19/USA/TX-HMH-739/2020\|EPI_ISL_546728\|2020-04 | ···················· | ···················· | ····················· | ···················· | ···················· | ···················· | |
| I | 126 | hCoV-19/USA/TX-HMH-7390/2020\|EPI_ISL_546729\|2020-0 | ···················· | ···················· | ····················· | ···················· | ···················· | ···················· | |
| I | 127 | hCoV-19/USA/TX-HMH-7391/2020\|EPI_ISL_546730\|2020-0 | ···················· | ····T··············· | ····················· | ···················· | ···················· | ···················· | |
| I | 128 | hCoV-19/USA/TX-HMH-7392/2020\|EPI_ISL_546731\|2020-0 | ···················· | ···················· | ····················· | ···················· | ···················· | ···················· | |
| I | 129 | hCoV-19/USA/TX-HMH-7395/2020\|EPI_ISL_546733\|2020-0 | ···················· | ···················· | ····················· | ···················· | ···················· | ···················· | |
| I | 130 | hCoV-19/USA/TX-HMH-7398/2020\|EPI_ISL_546736\|2020-0 | ···················· | ···················· | ····················· | ···················· | ···················· | ···················· | |
| I | 131 | hCoV-19/USA/TX-HMH-740/2020\|EPI_ISL_546738\|2020-04 | ···················· | ···················· | ····················· | ···················· | ···················· | ···················· | |
| I | 133 | hCoV-19/USA/TX-HMH-7401/2020\|EPI_ISL_546740\|2020-0 | ···················· | ···················· | ····················· | ···················· | ···················· | ···················· | |
| I | 134 | hCoV-19/USA/TX-HMH-7402/2020\|EPI_ISL_546741\|2020-0 | ···················· | ···················· | ····················· | ···················· | ···················· | ···················· | |
| I | 135 | hCoV-19/USA/TX-HMH-7403/2020\|EPI_ISL_546742\|2020-0 | ···················· | ···················· | ····················· | ···················· | ···················· | ···················· | |
| I | 136 | hCoV-19/USA/TX-HMH-7404/2020\|EPI_ISL_546743\|2020-0 | ···················· | ···················· | ····················· | ···················· | ···················· | ···················· | |
| I | 137 | hCoV-19/USA/TX-HMH-7406/2020\|EPI_ISL_546744\|2020-0 | ···················· | ·············A······ | ····················· | ···················· | ···················· | ···················· | |
| I | 140 | hCoV-19/USA/TX-HMH-741/2020\|EPI_ISL_546747\|2020-04 | ···················· | ···················· | ····················· | ···················· | ···················· | ···················· | |
| I | 141 | hCoV-19/USA/TX-HMH-7411/2020\|EPI_ISL_546749\|2020-0 | ···················· | ·············A······ | ····················· | ···················· | ···················· | ···················· | |
| I | 142 | hCoV-19/USA/TX-HMH-7412/2020\|EPI_ISL_546750\|2020-0 | ···················· | ···················· | ····················· | ···················· | ···················· | ···················· | |
| I | 144 | hCoV-19/USA/TX-HMH-742/2020\|EPI_ISL_546753\|2020-04 | ···················· | ···················· | ····················· | ···················· | ···················· | ···················· | |
| I | 145 | hCoV-19/USA/TX-HMH-7420/2020\|EPI_ISL_546754\|2020-0 | ···················· | ···················· | ····················· | ···················· | ···················· | ···················· | |
| I | 150 | hCoV-19/USA/TX-HMH-7479/2020\|EPI_ISL_546790\|2020-0 | ···················· | ···················· | ····················· | ···················· | ···················· | ···················· | |
| I | 151 | hCoV-19/USA/TX-HMH-7481/2020\|EPI_ISL_546792\|2020-0 | ···················· | ···················· | ····················· | ···················· | ···················· | ···················· | |
| I | 152 | hCoV-19/USA/TX-HMH-7482/2020\|EPI_ISL_546793\|2020-0 | ···················· | ···················· | ····················· | ···················· | ···················· | ···················· | |
| I | 153 | hCoV-19/USA/TX-HMH-7484/2020\|EPI_ISL_546794\|2020-0 | ···················· | ·············A······ | ····················· | ···················· | ···················· | ···················· | |
| I | 154 | hCoV-19/USA/TX-HMH-7487/2020\|EPI_ISL_546795\|2020-0 | ···················· | ···················· | ····················· | ···················· | ···················· | ···················· | |
| I | 155 | hCoV-19/USA/TX-HMH-749/2020\|EPI_ISL_546797\|2020-04 | ···················· | ···················· | ····················· | ···················· | ···················· | ···················· | |
| I | 156 | hCoV-19/USA/TX-HMH-6058/2020\|EPI_ISL_547200\|2020-0 | ···················· | ···················· | ····················· | ···················· | ···················· | ···················· | |
| I | 157 | hCoV-19/USA/TX-HMH-606/2020\|EPI_ISL_547201\|2020-04 | ···················· | ···················· | ····················· | ···················· | ···················· | ···················· | |
| I | 158 | hCoV-19/USA/TX-HMH-6060/2020\|EPI_ISL_547202\|2020-0 | ···················· | ···················· | ····················· | ···················· | ···················· | ···················· | |
| I | 160 | hCoV-19/USA/TX-HMH-6062/2020\|EPI_ISL_547204\|2020-0 | ···················· | ···················· | ····················· | ···················· | ···················· | ···················· | |
| I | 161 | hCoV-19/USA/TX-HMH-6063/2020\|EPI_ISL_547205\|2020-0 | ···················· | ···················· | ····················· | ···················· | ···················· | ···················· | |
| I | 165 | hCoV-19/USA/TX-HMH-6067/2020\|EPI_ISL_547209\|2020-0 | ···················· | ·············A······ | ····················· | ···················· | ···················· | ···················· | |
| I | 169 | hCoV-19/USA/TX-HMH-6072/2020\|EPI_ISL_547213\|2020-0 | ···················· | ·A·················· | ····················· | ···················· | ···················· | ···················· | |
| I | 171 | hCoV-19/USA/TX-HMH-6078/2020\|EPI_ISL_547215\|2020-0 | ···················· | ···················· | ····················· | ···················· | ···················· | NN·················· | |
| I | 172 | hCoV-19/USA/TX-HMH-6079/2020\|EPI_ISL_547216\|2020-0 | ···················· | ·············A······ | ····················· | ···················· | ··············N····· | NN·N················ | 1 |
| I | 173 | hCoV-19/USA/TX-HMH-608/2020\|EPI_ISL_547217\|2020-04 | ···················· | ···················· | ····················· | ···················· | ···················· | ···················· | |
| I | 177 | hCoV-19/USA/TX-HMH-6084/2020\|EPI_ISL_547222\|2020-0 | ···················· | ···················· | ····················· | ···················· | ···················· | ···················· | |
| I | 182 | hCoV-19/USA/TX-HMH-609/2020\|EPI_ISL_547228\|2020-04 | ···················· | ···················· | ····················· | ···················· | ···················· | ···················· | |
| I | 187 | hCoV-19/USA/TX-HMH-6099/2020\|EPI_ISL_547236\|2020-0 | ···················· | ·············A······ | ····················· | ···················· | ··············N····· | ···················· | |
| I | 188 | hCoV-19/USA/TX-HMH-6100/2020\|EPI_ISL_547237\|2020-0 | ···················· | ···················· | ····················· | ···················· | ···················· | ···················· | |
| I | 191 | hCoV-19/USA/CA-CZB-3884/2020\|EPI_ISL_548500\|2020-0 | ···················· | ···················· | ····················· | ···················· | ···················· | ···················· | |
| I | 192 | hCoV-19/USA/CA-CZB-3944/2020\|EPI_ISL_548501\|2020-0 | ···················· | ···················· | ····················· | ···················· | ···················· | ···················· | |
| I | 193 | hCoV-19/USA/CA-CZB-3904/2020\|EPI_ISL_548502\|2020-0 | ···················· | ···················· | ····················· | ···················· | ···················· | ···················· | |
| I | 194 | hCoV-19/USA/CA-CZB-3908/2020\|EPI_ISL_548504\|2020-0 | ···················· | ···················· | ····················· | ···················· | ···················· | ···················· | |
| I | 195 | hCoV-19/USA/CA-CZB-3953/2020\|EPI_ISL_548505\|2020-0 | ···················· | ···················· | ····················· | ···················· | ···················· | ···················· | |
| I | 196 | hCoV-19/USA/CA-CZB-3938/2020\|EPI_ISL_548506\|2020-0 | ···················· | ···················· | ····················· | ···················· | ···················· | ···················· | |
| I | 197 | hCoV-19/USA/CA-CZB-3942/2020\|EPI_ISL_548507\|2020-0 | ···················· | ···················· | ····················· | ···················· | ···················· | ···················· | |
| I | 198 | hCoV-19/USA/CA-CZB-3931/2020\|EPI_ISL_548508\|2020-0 | ···················· | ···················· | ····················· | ···················· | ···················· | ···················· | |
| I | 199 | hCoV-19/USA/CA-CZB-3873/2020\|EPI_ISL_548509\|2020-0 | ···················· | ···················· | ····················· | ···················· | ···················· | ···················· | |

```
I 200 hCoV-19/USA/CA-CZB-3933/2020|EPI_ISL_548510|2020-0 ···················· ·A·················· ···················· ···················· ···················· ····················
I 201 hCoV-19/USA/CA-CZB-3874/2020|EPI_ISL_548511|2020-0 ···················· ·A·················· ···················· ···················· ···················· ····················
I 202 hCoV-19/USA/CA-CZB-3886/2020|EPI_ISL_548512|2020-0 ···················· ···················· ···················· ···················· ···················· ····················
I 203 hCoV-19/USA/CA-CZB-3951/2020|EPI_ISL_548513|2020-0 ···················· ···················· ···················· ···················· ···················· ····················
I 204 hCoV-19/USA/CA-CZB-3875/2020|EPI_ISL_548514|2020-0 ···················· ·A·················· ···················· ···················· ···················· ····················
I 205 hCoV-19/USA/CA-CZB-3935/2020|EPI_ISL_548515|2020-0 ···················· ···················· ···················· ···················· ···················· ····················
I 206 hCoV-19/USA/CA-CZB-3929/2020|EPI_ISL_548516|2020-0 ···················· ···················· ···················· ···················· ···················· ····················
I 207 hCoV-19/USA/CA-CZB-3900/2020|EPI_ISL_548517|2020-0 ···················· ···················· ···················· ···················· ···················· ····················
I 208 hCoV-19/USA/CA-CZB-3883/2020|EPI_ISL_548518|2020-0 ···················· ·A·················· ···················· ···················· ···················· ····················
I 209 hCoV-19/USA/CA-CZB-3899/2020|EPI_ISL_548519|2020-0 ···················· ···················· ···················· ···················· ···················· ····················
I 210 hCoV-19/USA/CA-CZB-3949/2020|EPI_ISL_548520|2020-0 ···················· ·A·················· ···················· ···················· ···················· ····················
I 211 hCoV-19/USA/CA-CZB-3902/2020|EPI_ISL_548521|2020-0 ···················· ···················· ···················· ···················· ···················· ····················
I 212 hCoV-19/USA/CA-CZB-3952/2020|EPI_ISL_548522|2020-0 ···················· ·A·················· ···················· ···················· ···················· ····················
I 213 hCoV-19/USA/CA-CZB-3922/2020|EPI_ISL_548524|2020-0 ···················· ···················· ···················· ···················· ···················· ····················
I 214 hCoV-19/USA/CA-CZB-3894/2020|EPI_ISL_548525|2020-0 ···················· ···················· ···················· ···················· ···················· ····················
I 215 hCoV-19/USA/CA-CZB-3914/2020|EPI_ISL_548527|2020-0 ···················· ···················· ···················· ···················· ···················· ····················
I 216 hCoV-19/USA/CA-CZB-3916/2020|EPI_ISL_548528|2020-0 ···················· ···················· ···················· ···················· ···················· ····················
I 217 hCoV-19/USA/CA-CZB-3869/2020|EPI_ISL_548529|2020-0 ···················· ·A·················· ···················· ···················· ···················· ····················
I 218 hCoV-19/USA/CA-CZB-3939/2020|EPI_ISL_548531|2020-0 ···················· ···················· ···················· ···················· ···················· ····················
I 219 hCoV-19/USA/CA-CZB-3906/2020|EPI_ISL_548532|2020-0 ···················· ···················· ···················· ···················· ···················· ····················
I 220 hCoV-19/USA/CA-CZB-3887/2020|EPI_ISL_548533|2020-0 ···················· ·A·················· ···················· ···················· ···················· ····················
I 221 hCoV-19/USA/CA-CZB-3930/2020|EPI_ISL_548534|2020-0 ···················· ···················· ···················· ···················· ···················· ····················
I 222 hCoV-19/USA/CA-CZB-3923/2020|EPI_ISL_548535|2020-0 ···················· ·A·················· ···················· ···················· ···················· ···A················
I 223 hCoV-19/USA/CA-CZB-3868/2020|EPI_ISL_548536|2020-0 ···················· ·A·················· ···················· ···················· ···················· ····················
I 224 hCoV-19/USA/CA-CZB-3925/2020|EPI_ISL_548537|2020-0 ···················· ·A·················· ···················· ···················· ···················· ····················
I 225 hCoV-19/USA/CA-CZB-3945/2020|EPI_ISL_548538|2020-0 ···················· ···················· ···················· ···················· ···················· ····················
I 226 hCoV-19/USA/CA-CZB-3911/2020|EPI_ISL_548539|2020-0 ···················· ·A·················· ···················· ···················· ···················· ····················
I 227 hCoV-19/USA/CA-CZB-3912/2020|EPI_ISL_548540|2020-0 ···················· ···················· ···················· ···················· ···················· ····················
I 228 hCoV-19/USA/CA-CZB-3896/2020|EPI_ISL_548541|2020-0 ···················· ·A·················· ···················· ···················· ···················· ····················
I 229 hCoV-19/USA/CA-CZB-3901/2020|EPI_ISL_548542|2020-0 ···················· ···················· ···················· ···················· ···················· ····················
I 230 hCoV-19/USA/CA-CZB-3890/2020|EPI_ISL_548543|2020-0 ···················· ·A·················· ···················· ···················· ···················· ····················
I 231 hCoV-19/USA/CA-CZB-3907/2020|EPI_ISL_548544|2020-0 ···················· ·A·················· ···················· ···················· ···················· ····················
I 232 hCoV-19/USA/CA-CZB-3909/2020|EPI_ISL_548545|2020-0 ···················· ·A·················· ···················· ···················· ···················· ····················
I 233 hCoV-19/USA/CA-CZB-3881/2020|EPI_ISL_548546|2020-0 ···················· ·A·················· ···················· ···················· ···················· ····················
I 234 hCoV-19/USA/CA-CZB-3893/2020|EPI_ISL_548547|2020-0 ···················· ·A·················· ···················· ···················· ···················· ····················
I 235 hCoV-19/USA/CA-CZB-3924/2020|EPI_ISL_548548|2020-0 ···················· ·A·················· ···················· ···················· ···················· ····················
I 236 hCoV-19/USA/CA-CZB-3927/2020|EPI_ISL_548549|2020-0 ···················· ·A·················· ···················· ···················· ···················· ····················
I 237 hCoV-19/USA/CA-CZB-3878/2020|EPI_ISL_548550|2020-0 ···················· ···················· ···················· ···················· ···················· ····················
I 238 hCoV-19/USA/CA-CZB-3880/2020|EPI_ISL_548551|2020-0 ···················· ·A·················· ···················· ···················· ···················· ····················
I 239 hCoV-19/USA/CA-CZB-3932/2020|EPI_ISL_548552|2020-0 ···················· ···················· ···················· ···················· ···················· ····················
I 240 hCoV-19/USA/CA-CZB-3876/2020|EPI_ISL_548553|2020-0 ···················· ·A·················· ···················· ···················· ···················· ····················
I 241 hCoV-19/USA/CA-CZB-3936/2020|EPI_ISL_548554|2020-0 ···················· ···················· ···················· ···················· ···················· ····················
I 242 hCoV-19/USA/CA-CZB-3955/2020|EPI_ISL_548555|2020-0 ···················· ···················· ···················· ···················· ···················· ····················
I 243 hCoV-19/USA/CA-CZB-3941/2020|EPI_ISL_548556|2020-0 ···················· ·A·················· ···················· ···················· ···················· ····················
I 244 hCoV-19/USA/CA-CZB-3918/2020|EPI_ISL_548557|2020-0 ···················· ···················· ···················· ···················· ···················· ····················
I 245 hCoV-19/USA/CA-CZB-3948/2020|EPI_ISL_548558|2020-0 ···················· ·A·················· ···················· ···················· ···················· ····················
I 246 hCoV-19/USA/CA-CZB-3872/2020|EPI_ISL_548559|2020-0 ···················· ···················· ···················· ···················· ···················· ····················
I 247 hCoV-19/USA/CA-CZB-3903/2020|EPI_ISL_548560|2020-0 ···················· ·A·················· ···················· ···················· ···················· ····················
I 248 hCoV-19/USA/CA-CZB-3913/2020|EPI_ISL_548561|2020-0 ···················· ·A·················· ···················· ···················· ···················· ····················
I 249 hCoV-19/USA/CA-CZB-3879/2020|EPI_ISL_548562|2020-0 ···················· ···················· ···················· ···················· ···················· ····················
I 250 hCoV-19/USA/CA-CZB-3867/2020|EPI_ISL_548563|2020-0 ···················· ·A·················· ···················· ···················· ···················· ····················
I 251 hCoV-19/USA/CA-CZB-3921/2020|EPI_ISL_548564|2020-0 ···················· ···················· ···················· ···················· ···················· ····················
I 252 hCoV-19/USA/CA-CZB-3947/2020|EPI_ISL_548565|2020-0 ···················· ···················· ···················· ···················· ···················· ····················
I 253 hCoV-19/USA/CA-CZB-3937/2020|EPI_ISL_548566|2020-0 ···················· ···················· ···················· ···················· ···················· ····················
I 254 hCoV-19/USA/CA-CZB-3910/2020|EPI_ISL_548567|2020-0 ···················· ·A·················· ···················· ···················· ···················· ····················
I 255 hCoV-19/USA/CA-CZB-3895/2020|EPI_ISL_548568|2020-0 ···················· ·A·················· ···················· ···················· ···················· ····················
I 256 hCoV-19/USA/CA-CZB-3943/2020|EPI_ISL_548569|2020-0 ···················· ·A··A··············· ···················· ···················· ···················· ····················
I 257 hCoV-19/USA/TX-HMH-2813/2020|EPI_ISL_543789|2020-0 ···················· ···················· ···················· ···················· ···················· ····················
I 258 hCoV-19/USA/TX-HMH-2814/2020|EPI_ISL_543790|2020-0 ···················· ···················· ···················· ···················· ···················· ····················
I 259 hCoV-19/USA/TX-HMH-2815/2020|EPI_ISL_543791|2020-0 ···················· ···················· ···················· ···················· ···················· ····················
I 260 hCoV-19/USA/TX-HMH-2816/2020|EPI_ISL_543792|2020-0 ···················· ···················· ···················· ···················· ···················· ····················
I 261 hCoV-19/USA/TX-HMH-2818/2020|EPI_ISL_543794|2020-0 ···················· ···················· ···················· ···················· ···················· ····················
I 262 hCoV-19/USA/TX-HMH-2819/2020|EPI_ISL_543795|2020-0 ···················· ···················· ···················· ···················· ···················· ····················
I 263 hCoV-19/USA/TX-HMH-2820/2020|EPI_ISL_543796|2020-0 ···················· ···················· ···················· ···················· ···················· ····················
I 264 hCoV-19/USA/TX-HMH-2821/2020|EPI_ISL_543797|2020-0 ···················· ···················· ···················· ···················· ···················· ····················
I 265 hCoV-19/USA/TX-HMH-3121/2020|EPI_ISL_544007|2020-0 ···················· ······A············· ···················· ···················· ···················· ····················
I 266 hCoV-19/USA/TX-HMH-3122/2020|EPI_ISL_544008|2020-0 ···················· ···················· ···················· ···················· ···················· ····················
I 267 hCoV-19/USA/TX-HMH-3123/2020|EPI_ISL_544009|2020-0 ···················· ···················· ···················· ···················· ···················· ····················
I 268 hCoV-19/USA/TX-HMH-3124/2020|EPI_ISL_544010|2020-0 ···················· ···················· ···················· ···················· ···················· ····················
I 269 hCoV-19/USA/TX-HMH-3126/2020|EPI_ISL_544011|2020-0 ···················· ······A············· ···················· ···················· ···················· ····················
I 270 hCoV-19/USA/TX-HMH-3127/2020|EPI_ISL_544012|2020-0 ···················· ···················· ···················· ···················· ···················· ····················
I 271 hCoV-19/USA/TX-HMH-3129/2020|EPI_ISL_544014|2020-0 ···················· ···················· ···················· ···················· ···················· ····················
I 272 hCoV-19/USA/TX-HMH-3133/2020|EPI_ISL_544016|2020-0 ···················· ···················· ···················· ···················· ···················· ····················
I 273 hCoV-19/USA/TX-HMH-3134/2020|EPI_ISL_544017|2020-0 ···················· ···················· ···················· ···················· ···················· ····················
I 274 hCoV-19/USA/TX-HMH-3136/2020|EPI_ISL_544018|2020-0 ···················· ······A············· ···················· ···················· ···················· ····················
I 275 hCoV-19/USA/TX-HMH-3137/2020|EPI_ISL_544019|2020-0 ···················· ···················· ···················· ···················· ···················· ····················
I 276 hCoV-19/USA/TX-HMH-3138/2020|EPI_ISL_544020|2020-0 ···················· ···················· ···················· ···················· ···················· ···A················
I 277 hCoV-19/USA/TX-HMH-3139/2020|EPI_ISL_544021|2020-0 ···················· ···················· ···················· ···················· ···················· ····················
I 278 hCoV-19/USA/TX-HMH-3143/2020|EPI_ISL_544022|2020-0 ···················· ···················· ···················· ···················· ···················· ····················
I 279 hCoV-19/USA/TX-HMH-3144/2020|EPI_ISL_544023|2020-0 ···················· ·············A······ ···················· ···················· ···················· ····················
```

| | | | | | | | |
|---|---|---|---|---|---|---|---|
| I | 280 hCoV-19/USA/TX-HMH-3145/2020\|EPI_ISL_544024\|2020-0 | ···················· | ···················· | ···················· | ···················· | ···················· | ···················· |
| I | 283 hCoV-19/USA/TX-HMH-3150/2020\|EPI_ISL_544028\|2020-0 | ···················· | ···················· | ···················· | ···················· | ···················· | ···················· |
| I | 284 hCoV-19/USA/TX-HMH-3152/2020\|EPI_ISL_544030\|2020-0 | ···················· | ···················· | ···················· | ···················· | ···················· | ···················· |
| I | 285 hCoV-19/USA/TX-HMH-3156/2020\|EPI_ISL_544032\|2020-0 | ···················· | ···················· | ···················· | ···················· | ···················· | ···················· |
| I | 286 hCoV-19/USA/TX-HMH-3157/2020\|EPI_ISL_544033\|2020-0 | ···················· | ···················· | ···················· | ···················· | ···················· | ···················· |
| I | 287 hCoV-19/USA/TX-HMH-3158/2020\|EPI_ISL_544034\|2020-0 | ···················· | ···················· | ···················· | ···················· | ···················· | ···················· |
| I | 288 hCoV-19/USA/TX-HMH-3159/2020\|EPI_ISL_544035\|2020-0 | ···················· | ···················· | ···················· | ···················· | ···················· | ···················· |
| I | 289 hCoV-19/USA/TX-HMH-3160/2020\|EPI_ISL_544036\|2020-0 | ···················· | ···················· | ···················· | ···················· | ···················· | ···················· |
| I | 290 hCoV-19/USA/TX-HMH-3161/2020\|EPI_ISL_544037\|2020-0 | ···················· | ···················· | ···················· | ···················· | ··············N····· | ···················· |
| I | 291 hCoV-19/USA/TX-HMH-3162/2020\|EPI_ISL_544038\|2020-0 | ···················· | ···················· | ···················· | ···················· | ···················· | ···················· |
| I | 292 hCoV-19/USA/TX-HMH-3163/2020\|EPI_ISL_544039\|2020-0 | ···················· | ···················· | ···················· | ···················· | ···················· | ···················· |
| I | 293 hCoV-19/USA/TX-HMH-3164/2020\|EPI_ISL_544040\|2020-0 | ···················· | ···················· | ···················· | ···················· | ···················· | ···················· |
| I | 294 hCoV-19/USA/TX-HMH-3165/2020\|EPI_ISL_544041\|2020-0 | ···················· | ···················· | ···················· | ···················· | ···················· | ···················· |
| I | 295 hCoV-19/USA/TX-HMH-3166/2020\|EPI_ISL_544042\|2020-0 | ···················· | ···················· | ···················· | ···················· | ···················· | ···················· |
| I | 296 hCoV-19/USA/TX-HMH-3282/2020\|EPI_ISL_544100\|2020-0 | ···················· | ···················· | ···················· | ···················· | ···················· | ···················· |
| I | 297 hCoV-19/USA/TX-HMH-3283/2020\|EPI_ISL_544101\|2020-0 | ···················· | ···················· | ···················· | ···················· | ···················· | ···················· |
| I | 298 hCoV-19/USA/TX-HMH-3284/2020\|EPI_ISL_544102\|2020-0 | ···················· | ···················· | ···················· | ···················· | ···················· | ···················· |
| I | 299 hCoV-19/USA/TX-HMH-3288/2020\|EPI_ISL_544105\|2020-0 | ···················· | ···················· | ···················· | ···················· | ···················· | ···················· |
| I | 300 hCoV-19/USA/TX-HMH-3289/2020\|EPI_ISL_544106\|2020-0 | ···················· | ···················· | ···················· | ···················· | ···················· | ···················· |
| I | 301 hCoV-19/USA/TX-HMH-3290/2020\|EPI_ISL_544107\|2020-0 | ···················· | ···················· | ···················· | ···················· | ···················· | ···················· |
| I | 303 hCoV-19/USA/TX-HMH-3293/2020\|EPI_ISL_544110\|2020-0 | ···················· | ···················· | ···················· | ···················· | ···················· | ···················· |
| I | 304 hCoV-19/USA/TX-HMH-3297/2020\|EPI_ISL_544112\|2020-0 | ···················· | ···················· | ···················· | ···················· | ···················· | ···················· |
| I | 305 hCoV-19/USA/TX-HMH-3300/2020\|EPI_ISL_544113\|2020-0 | ···················· | ···················· | ···················· | ···················· | ···················· | ···················· |
| I | 306 hCoV-19/USA/TX-HMH-3301/2020\|EPI_ISL_544114\|2020-0 | ···················· | ···················· | ···················· | ···················· | ···················· | ···················· |
| I | 307 hCoV-19/USA/TX-HMH-3302/2020\|EPI_ISL_544115\|2020-0 | ···················· | ······A············· | ···················· | ···················· | ···················· | ···················· |
| I | 308 hCoV-19/USA/TX-HMH-3304/2020\|EPI_ISL_544117\|2020-0 | ···················· | ···················· | ···················· | ···················· | ···················· | ···················· |
| I | 309 hCoV-19/USA/TX-HMH-3306/2020\|EPI_ISL_544118\|2020-0 | ···················· | ···················· | ···················· | ···················· | ···················· | ···················· |
| I | 310 hCoV-19/USA/TX-HMH-3308/2020\|EPI_ISL_544120\|2020-0 | ···················· | ···················· | ···················· | ···················· | ···················· | ···················· |
| I | 311 hCoV-19/USA/TX-HMH-3309/2020\|EPI_ISL_544121\|2020-0 | ···················· | ···················· | ···················· | ···················· | ···················· | ···················· |
| I | 313 hCoV-19/USA/TX-HMH-3311/2020\|EPI_ISL_544123\|2020-0 | ···················· | ···················· | ···················· | ···················· | ···················· | ···················· |
| I | 314 hCoV-19/USA/TX-HMH-3314/2020\|EPI_ISL_544124\|2020-0 | ···················· | ···················· | ···················· | ···················· | ···················· | ···················· |
| I | 315 hCoV-19/USA/TX-HMH-3316/2020\|EPI_ISL_544126\|2020-0 | ···················· | ···················· | ···················· | ···················· | ···················· | ···················· |
| I | 316 hCoV-19/USA/TX-HMH-3317/2020\|EPI_ISL_544127\|2020-0 | ···················· | ···················· | ···················· | ···················· | ···················· | ···················· |
| I | 317 hCoV-19/USA/TX-HMH-2491/2020\|EPI_ISL_545338\|2020-0 | ···················· | ···················· | ···················· | ···················· | ···················· | ···················· |
| I | 318 hCoV-19/USA/TX-HMH-2494/2020\|EPI_ISL_545339\|2020-0 | ···················· | ···················· | ···················· | ···················· | ···················· | ···················· |
| I | 319 hCoV-19/USA/TX-HMH-2495/2020\|EPI_ISL_545340\|2020-0 | ···················· | ···················· | ···················· | ···················· | ···················· | ···················· |
| I | 320 hCoV-19/USA/TX-HMH-2503/2020\|EPI_ISL_545342\|2020-0 | ···················· | ···················· | ···················· | ···················· | ···················· | ···················· |
| I | 321 hCoV-19/USA/TX-HMH-2505/2020\|EPI_ISL_545343\|2020-0 | ···················· | ···················· | ···················· | ···················· | ···················· | ···················· |
| I | 322 hCoV-19/USA/TX-HMH-2506/2020\|EPI_ISL_545344\|2020-0 | ···················· | ···················· | ···················· | ···················· | ···················· | ···················· |
| I | 323 hCoV-19/USA/TX-HMH-2507/2020\|EPI_ISL_545345\|2020-0 | ···················· | ···················· | ···················· | ···················· | ···················· | ···················· |
| I | 324 hCoV-19/USA/TX-HMH-2509/2020\|EPI_ISL_545346\|2020-0 | ···················· | ···················· | ···················· | ···················· | ···················· | ···················· |
| I | 325 hCoV-19/USA/TX-HMH-2510/2020\|EPI_ISL_545347\|2020-0 | ···················· | ···················· | ···················· | ···················· | ···················· | ···················· |
| I | 326 hCoV-19/USA/TX-HMH-2512/2020\|EPI_ISL_545348\|2020-0 | ···················· | ··············C····· | ···················· | ···················· | ···················· | ···················· |
| I | 327 hCoV-19/USA/TX-HMH-2513/2020\|EPI_ISL_545349\|2020-0 | ···················· | ···················· | ···················· | ···················· | ···················· | ···················· |
| I | 328 hCoV-19/USA/TX-HMH-2515/2020\|EPI_ISL_545351\|2020-0 | ···················· | ···················· | ···················· | ···················· | ···················· | ···················· |
| I | 329 hCoV-19/USA/TX-HMH-2516/2020\|EPI_ISL_545352\|2020-0 | ···················· | ···················· | ···················· | ···················· | ···················· | ···················· |
| I | 330 hCoV-19/USA/TX-HMH-2517/2020\|EPI_ISL_545353\|2020-0 | ···················· | ···················· | ···················· | ···················· | ···················· | ···················· |
| I | 331 hCoV-19/USA/TX-HMH-2518/2020\|EPI_ISL_545354\|2020-0 | ···················· | ···················· | ···················· | ···················· | ···················· | ···················· |
| I | 332 hCoV-19/USA/TX-HMH-2519/2020\|EPI_ISL_545355\|2020-0 | ···················· | ···················· | ···················· | ···················· | ···················· | ···················· |
| I | 333 hCoV-19/USA/TX-HMH-2520/2020\|EPI_ISL_545356\|2020-0 | ···················· | ···················· | ···················· | ···················· | ···················· | ···················· |
| I | 334 hCoV-19/USA/TX-HMH-2521/2020\|EPI_ISL_545357\|2020-0 | ···················· | ···················· | ···················· | ···················· | ···················· | ···················· |
| I | 335 hCoV-19/USA/TX-HMH-2522/2020\|EPI_ISL_545358\|2020-0 | ···················· | ···················· | ···················· | ···················· | ···················· | ···················· |
| I | 336 hCoV-19/USA/TX-HMH-2523/2020\|EPI_ISL_545359\|2020-0 | ···················· | ···················· | ···················· | ···················· | ···················· | ···················· |
| I | 337 hCoV-19/USA/TX-HMH-2524/2020\|EPI_ISL_545360\|2020-0 | ···················· | ···················· | ···················· | ···················· | ···················· | ···················· |
| I | 338 hCoV-19/USA/TX-HMH-2525/2020\|EPI_ISL_545361\|2020-0 | ···················· | ···················· | ···················· | ···················· | ···················· | ···················· |
| I | 339 hCoV-19/USA/TX-HMH-2526/2020\|EPI_ISL_545362\|2020-0 | ···················· | ···················· | ···················· | ···················· | ···················· | ···················· |
| I | 340 hCoV-19/USA/TX-HMH-2527/2020\|EPI_ISL_545363\|2020-0 | ···················· | ···················· | ···················· | ···················· | ···················· | ···················· |
| I | 341 hCoV-19/USA/TX-HMH-2532/2020\|EPI_ISL_545364\|2020-0 | ···················· | ···················· | ···················· | ···················· | ···················· | ···················· |
| I | 342 hCoV-19/USA/TX-HMH-2534/2020\|EPI_ISL_545366\|2020-0 | ···················· | ···················· | ···················· | ···················· | ···················· | ···················· |
| I | 343 hCoV-19/USA/TX-HMH-2536/2020\|EPI_ISL_545367\|2020-0 | ···················· | ···················· | ···················· | ···················· | ···················· | ···················· |
| I | 344 hCoV-19/USA/TX-HMH-2537/2020\|EPI_ISL_545368\|2020-0 | ···················· | ···················· | ···················· | ···················· | ···················· | ···················· |
| I | 345 hCoV-19/USA/TX-HMH-2538/2020\|EPI_ISL_545369\|2020-0 | ···················· | ···················· | ···················· | ···················· | ···················· | ···················· |
| I | 346 hCoV-19/USA/TX-HMH-2540/2020\|EPI_ISL_545371\|2020-0 | ···················· | ···················· | ···················· | ···················· | ···················· | ···················· |
| I | 347 hCoV-19/USA/TX-HMH-2541/2020\|EPI_ISL_545372\|2020-0 | ···················· | ···················· | ···················· | ···················· | ···················· | ···················· |
| I | 348 hCoV-19/USA/TX-HMH-2543/2020\|EPI_ISL_545373\|2020-0 | ···················· | ···················· | ···················· | ···················· | ···················· | ···················· |
| I | 349 hCoV-19/USA/TX-HMH-2544/2020\|EPI_ISL_545374\|2020-0 | ···················· | ···················· | ···················· | ···················· | ···················· | ···················· |
| I | 350 hCoV-19/USA/TX-HMH-2610/2020\|EPI_ISL_545426\|2020-0 | ···················· | ···················· | ···················· | ···················· | ···················· | ···················· |
| I | 351 hCoV-19/USA/TX-HMH-2612/2020\|EPI_ISL_545427\|2020-0 | ···················· | ···················· | ···················· | ···················· | ···················· | ···················· |
| I | 352 hCoV-19/USA/TX-HMH-2615/2020\|EPI_ISL_545429\|2020-0 | ···················· | ···················· | ···················· | ···················· | ···················· | ···················· |
| I | 353 hCoV-19/USA/TX-HMH-2618/2020\|EPI_ISL_545431\|2020-0 | ···················· | ···················· | ···················· | ···················· | ···················· | ···················· |
| I | 354 hCoV-19/USA/TX-HMH-2620/2020\|EPI_ISL_545432\|2020-0 | ···················· | ···················· | ···················· | ···················· | ···················· | ···················· |
| I | 356 hCoV-19/USA/TX-HMH-2626/2020\|EPI_ISL_545435\|2020-0 | ···················· | ······A············· | ···················· | ···················· | ···················· | ···················· |
| I | 357 hCoV-19/USA/TX-HMH-2628/2020\|EPI_ISL_545436\|2020-0 | ···················· | ···················· | ···················· | ···················· | ···················· | ···················· |
| I | 358 hCoV-19/USA/TX-HMH-2629/2020\|EPI_ISL_545437\|2020-0 | ···················· | ···················· | ···················· | ···················· | ···················· | ···················· |
| I | 359 hCoV-19/USA/TX-HMH-2631/2020\|EPI_ISL_545438\|2020-0 | ···················· | ···················· | ···················· | ···················· | ···················· | ···················· |
| I | 360 hCoV-19/USA/TX-HMH-2632/2020\|EPI_ISL_545439\|2020-0 | ···················· | ···················· | ···················· | ···················· | ···················· | ···················· |
| I | 361 hCoV-19/USA/TX-HMH-2633/2020\|EPI_ISL_545440\|2020-0 | ···················· | ···················· | ···················· | ···················· | ···················· | ···················· |
| I | 362 hCoV-19/USA/TX-HMH-2634/2020\|EPI_ISL_545441\|2020-0 | ···················· | ···················· | ···················· | ···················· | ···················· | ···················· |
| I | 363 hCoV-19/USA/TX-HMH-2635/2020\|EPI_ISL_545442\|2020-0 | ···················· | ···················· | ···················· | ···················· | ···················· | ···················· |
| I | 364 hCoV-19/USA/TX-HMH-2636/2020\|EPI_ISL_545443\|2020-0 | ···················· | ···················· | ···················· | ···················· | ···················· | ···················· |

| | | | | | | | | |
|---|---|---|---|---|---|---|---|---|
| I | 365 hCoV-19/USA/TX-HMH-2637/2020\|EPI_ISL_545444\|2020-0 | ···················· | ···················· | ···················· | ···················· | ···················· | ···················· | |
| I | 366 hCoV-19/USA/TX-HMH-2638/2020\|EPI_ISL_545445\|2020-0 | ···················· | ···················· | ···················· | ···················· | ···················· | ···················· | |
| I | 367 hCoV-19/USA/TX-HMH-2639/2020\|EPI_ISL_545446\|2020-0 | ···················· | ···················· | ···················· | ···················· | ···················· | ···················· | |
| I | 368 hCoV-19/USA/TX-HMH-2642/2020\|EPI_ISL_545448\|2020-0 | ···················· | ···················· | ···················· | ···················· | ···················· | ···················· | |
| I | 369 hCoV-19/USA/TX-HMH-2644/2020\|EPI_ISL_545449\|2020-0 | ···················· | ···················· | ···················· | ···················· | ···················· | ···················· | |
| I | 370 hCoV-19/USA/TX-HMH-2649/2020\|EPI_ISL_545450\|2020-0 | ···················· | ···················· | ···················· | ···················· | ···················· | ···················· | |
| I | 371 hCoV-19/USA/TX-HMH-2650/2020\|EPI_ISL_545451\|2020-0 | ···················· | ···················· | ···················· | ···················· | ···················· | ···················· | |
| I | 372 hCoV-19/USA/TX-HMH-2655/2020\|EPI_ISL_545453\|2020-0 | ···················· | ···················· | ···················· | ···················· | ···················· | ···················· | |
| I | 373 hCoV-19/USA/TX-HMH-2656/2020\|EPI_ISL_545454\|2020-0 | ···················· | ···················· | ···················· | ···················· | ···················· | ···················· | |
| I | 374 hCoV-19/USA/TX-HMH-2657/2020\|EPI_ISL_545455\|2020-0 | ···················· | ···················· | ···················· | ···················· | ···················· | ···················· | |
| I | 376 hCoV-19/USA/TX-HMH-2659/2020\|EPI_ISL_545457\|2020-0 | ···················· | ···················· | ···················· | ···················· | ···················· | ···················· | |
| I | 377 hCoV-19/USA/TX-HMH-2660/2020\|EPI_ISL_545458\|2020-0 | ···················· | ···················· | ···················· | ···················· | ···················· | ···················· | |
| I | 378 hCoV-19/USA/TX-HMH-7312/2020\|EPI_ISL_546669\|2020-0 | ···················· | ···················· | ···················· | ···················· | ···················· | ···················· | |
| I | 379 hCoV-19/USA/TX-HMH-7313/2020\|EPI_ISL_546670\|2020-0 | ···················· | ···················· | ···················· | ···················· | ···················· | ···················· | |
| I | 380 hCoV-19/USA/TX-HMH-7314/2020\|EPI_ISL_546671\|2020-0 | ···················· | ···················· | ···················· | ···················· | ···················· | ···················· | |
| I | 381 hCoV-19/USA/TX-HMH-7317/2020\|EPI_ISL_546672\|2020-0 | ···················· | ···················· | ···················· | ···················· | ···················· | ···················· | |
| I | 383 hCoV-19/USA/TX-HMH-7323/2020\|EPI_ISL_546675\|2020-0 | ···················· | ···················· | ···················· | ···················· | ···················· | ···················· | |
| I | 385 hCoV-19/USA/TX-HMH-7329/2020\|EPI_ISL_546677\|2020-0 | ···················· | ···················· | ···················· | ···················· | ···················· | ···················· | |
| I | 386 hCoV-19/USA/TX-HMH-733/2020\|EPI_ISL_546678\|2020-04 | ···················· | ···················· | ···················· | ···················· | ···················· | ···················· | |
| I | 388 hCoV-19/USA/TX-HMH-7331/2020\|EPI_ISL_546680\|2020-0 | ···················· | ············A······· | ···················· | ···················· | ···················· | ···················· | |
| I | 389 hCoV-19/USA/TX-HMH-7332/2020\|EPI_ISL_546681\|2020-0 | ···················· | ···················· | ···················· | ···················· | ···················· | ···················· | |
| I | 390 hCoV-19/USA/TX-HMH-7336/2020\|EPI_ISL_546683\|2020-0 | ···················· | ···················· | ···················· | ···················· | ···················· | ···················· | |
| I | 393 hCoV-19/USA/TX-HMH-7340/2020\|EPI_ISL_546687\|2020-0 | ···················· | ···················· | ···················· | ···················· | ···················· | ···················· | |
| I | 394 hCoV-19/USA/TX-HMH-7341/2020\|EPI_ISL_546688\|2020-0 | ···················· | ···················· | ···················· | ···················· | ···················· | ···A················ | |
| I | 395 hCoV-19/USA/TX-HMH-7342/2020\|EPI_ISL_546689\|2020-0 | ···················· | ···················· | ···················· | ···················· | ···················· | ···················· | |
| I | 397 hCoV-19/USA/TX-HMH-7346/2020\|EPI_ISL_546691\|2020-0 | ···················· | ···················· | ···················· | ···················· | ··············N····· | ···················· | |
| I | 398 hCoV-19/USA/TX-HMH-7347/2020\|EPI_ISL_546692\|2020-0 | ···················· | ············A······· | ···················· | ···················· | ···················· | ···················· | |
| I | 401 hCoV-19/USA/TX-HMH-7352/2020\|EPI_ISL_546697\|2020-0 | ···················· | ···················· | ···················· | ···················· | ··············NN···· | ···A··NN············ | 1 |
| I | 411 hCoV-19/USA/TX-HMH-7443/2020\|EPI_ISL_546771\|2020-0 | ···················· | ···················· | ···················· | ···················· | ···················· | ···················· | |
| I | 413 hCoV-19/USA/TX-HMH-745/2020\|EPI_ISL_546775\|2020-04 | ···················· | ···················· | ···················· | ···················· | ···················· | ···················· | |
| I | 423 hCoV-19/USA/TX-HMH-747/2020\|EPI_ISL_546786\|2020-04 | ···················· | ···················· | ···················· | ···················· | ···················· | ···················· | |
| I | 433 hCoV-19/USA/TX-HMH-7507/2020\|EPI_ISL_546808\|2020-0 | ···················· | ···················· | ···················· | ···················· | ···················· | ···················· | |
| I | 434 hCoV-19/USA/TX-HMH-751/2020\|EPI_ISL_546810\|2020-04 | ···················· | ···················· | ···················· | ···················· | ···················· | ···················· | |
| I | 435 hCoV-19/USA/TX-HMH-7510/2020\|EPI_ISL_546811\|2020-0 | ···················· | ···················· | ···················· | ···················· | ···················· | ···················· | |
| I | 437 hCoV-19/USA/TX-HMH-7515/2020\|EPI_ISL_546815\|2020-0 | ···················· | ···················· | ···················· | ···················· | ···················· | ···················· | |
| I | 438 hCoV-19/USA/TX-HMH-7516/2020\|EPI_ISL_546816\|2020-0 | ···················· | ···················· | ···················· | ···················· | ···················· | ···················· | |
| I | 439 hCoV-19/USA/TX-HMH-7522/2020\|EPI_ISL_546819\|2020-0 | ···················· | ···················· | ···················· | ···················· | ···················· | ···················· | |
| I | 443 hCoV-19/USA/TX-HMH-611/2020\|EPI_ISL_547245\|2020-04 | ···················· | ···················· | ···················· | ···················· | ···················· | ···················· | |
| I | 451 hCoV-19/USA/TX-HMH-612/2020\|EPI_ISL_547255\|2020-04 | ···················· | ···················· | ···················· | ···················· | ···················· | ···················· | |
| I | 455 hCoV-19/USA/TX-HMH-6129/2020\|EPI_ISL_547262\|2020-0 | ···················· | ············A······· | ···················· | ···················· | ···················· | ···N················ | |
| I | 460 hCoV-19/USA/TX-HMH-6147/2020\|EPI_ISL_547269\|2020-0 | ···················· | ···················· | ···················· | ···················· | ···················· | ···················· | |
| I | 470 hCoV-19/USA/TX-HMH-6159/2020\|EPI_ISL_547280\|2020-0 | ···················· | ···················· | ···················· | ···················· | ···················· | ···················· | |
| I | 471 hCoV-19/USA/TX-HMH-6163/2020\|EPI_ISL_547283\|2020-0 | ···················· | ············A······· | ···················· | ···················· | ···················· | ···················· | |
| I | 472 hCoV-19/USA/TX-HMH-6167/2020\|EPI_ISL_547286\|2020-0 | ···················· | ···················· | ···················· | ···················· | ···················· | ···················· | |
| I | 473 hCoV-19/USA/TX-HMH-6169/2020\|EPI_ISL_547287\|2020-0 | ···················· | ···················· | ···················· | ···················· | ···················· | ···················· | |
| I | 474 hCoV-19/USA/TX-HMH-617/2020\|EPI_ISL_547288\|2020-04 | ···················· | ···················· | ···················· | ···················· | ···················· | ···················· | |
| I | 475 hCoV-19/USA/TX-HMH-6171/2020\|EPI_ISL_547289\|2020-0 | ···················· | ···················· | ···················· | ···················· | ··············N····· | ········N··········· | |
| I | 477 hCoV-19/USA/TX-HMH-6176/2020\|EPI_ISL_547293\|2020-0 | ···················· | ···················· | ···················· | ···················· | ··············N····· | ···················· | |
| I | 478 hCoV-19/USA/TX-HMH-6178/2020\|EPI_ISL_547294\|2020-0 | ···················· | ············A······· | ···················· | ···················· | ···················· | ···················· | |
| I | 479 hCoV-19/USA/TX-HMH-6181/2020\|EPI_ISL_547295\|2020-0 | ···················· | ···················· | ···················· | ···················· | ···················· | ···················· | |
| I | 480 hCoV-19/USA/TX-HMH-6183/2020\|EPI_ISL_547296\|2020-0 | ···················· | ···················· | ···················· | ···················· | ···················· | ···················· | |
| I | 481 hCoV-19/USA/TX-HMH-6185/2020\|EPI_ISL_547297\|2020-0 | ···················· | ···················· | ···················· | ···················· | ···················· | ···················· | |
| I | 482 hCoV-19/USA/TX-HMH-6187/2020\|EPI_ISL_547298\|2020-0 | ···················· | ············A······· | ···················· | ···················· | ···················· | ···················· | |
| I | 483 hCoV-19/USA/TX-HMH-6188/2020\|EPI_ISL_547299\|2020-0 | ···················· | ···················· | ···················· | ···················· | ···················· | ···················· | |
| I | 484 hCoV-19/USA/CA-CZB-3696/2020\|EPI_ISL_548401\|2020-0 | ···················· | ···················· | ···················· | ···················· | ···················· | ···················· | |
| I | 485 hCoV-19/USA/CA-CZB-3871/2020\|EPI_ISL_548570\|2020-0 | ···················· | ·A·················· | ···················· | ···················· | ···················· | ···················· | |
| I | 486 hCoV-19/USA/CA-CZB-3954/2020\|EPI_ISL_548571\|2020-0 | ···················· | ·A·················· | ···················· | ···················· | ···················· | ···················· | |
| I | 487 hCoV-19/USA/CA-CZB-3917/2020\|EPI_ISL_548572\|2020-0 | ···················· | ·A·················· | ···················· | ···················· | ···················· | ···················· | |
| I | 488 hCoV-19/USA/CA-CZB-3940/2020\|EPI_ISL_548573\|2020-0 | ···················· | ·A·················· | ···················· | ···················· | ···················· | ···················· | |
| I | 489 hCoV-19/USA/CA-CZB-3934/2020\|EPI_ISL_548574\|2020-0 | ···················· | ·A·················· | ···················· | ···················· | ···················· | ···················· | |
| I | 490 hCoV-19/USA/CA-CZB-3920/2020\|EPI_ISL_548575\|2020-0 | ···················· | ···················· | ···················· | ···················· | ···················· | ···················· | |
| I | 491 hCoV-19/USA/CA-CZB-3870/2020\|EPI_ISL_548576\|2020-0 | ···················· | ·A·················· | ···················· | ···················· | ···················· | ···················· | |
| I | 492 hCoV-19/USA/CA-CZB-3950/2020\|EPI_ISL_548577\|2020-0 | ···················· | ·A·················· | ···················· | ···················· | ···················· | ···················· | |
| I | 493 hCoV-19/USA/CA-CZB-3885/2020\|EPI_ISL_548578\|2020-0 | ···················· | ·A·················· | ···················· | ···················· | ···················· | ···················· | |
| I | 494 hCoV-19/USA/CA-CZB-3915/2020\|EPI_ISL_548579\|2020-0 | ···················· | ···················· | ···················· | ···················· | ···················· | ···················· | |
| I | 495 hCoV-19/USA/CA-CZB-4063/2020\|EPI_ISL_548580\|2020-0 | ···················· | ···················· | ···················· | ···················· | ···················· | ···················· | |
| I | 496 hCoV-19/USA/CA-CZB-4072/2020\|EPI_ISL_548581\|2020-0 | ···················· | ···················· | ···················· | ···················· | ···················· | ···················· | |
| I | 497 hCoV-19/USA/CA-CZB-4029/2020\|EPI_ISL_548582\|2020-0 | ···················· | ···················· | ···················· | ···················· | ···················· | ···················· | |
| I | 498 hCoV-19/USA/CA-CZB-4044/2020\|EPI_ISL_548583\|2020-0 | ···················· | ·A·················· | ···················· | ···················· | ···················· | ···················· | |
| I | 499 hCoV-19/USA/CA-CZB-4016/2020\|EPI_ISL_548584\|2020-0 | ···················· | ·A·················· | ···················· | ···················· | ···················· | ···················· | |
| I | 500 hCoV-19/USA/CA-CZB-4076/2020\|EPI_ISL_548585\|2020-0 | ···················· | ···················· | ···················· | ···················· | ···················· | ···················· | |
| I | 501 hCoV-19/USA/CA-CZB-4061/2020\|EPI_ISL_548586\|2020-0 | ···················· | ···················· | ···················· | ···················· | ···················· | ···················· | |
| I | 502 hCoV-19/USA/CA-CZB-4017/2020\|EPI_ISL_548588\|2020-0 | ···················· | ·A·················· | ···················· | ···················· | ···················· | ···················· | |
| I | 503 hCoV-19/USA/CA-CZB-4100/2020\|EPI_ISL_548590\|2020-0 | ···················· | ···················· | ···················· | ···················· | ···················· | ···················· | |
| I | 504 hCoV-19/USA/CA-CZB-4082/2020\|EPI_ISL_548591\|2020-0 | ···················· | ···················· | ···················· | ···················· | ···················· | ···················· | |
| I | 505 hCoV-19/USA/CA-CZB-4058/2020\|EPI_ISL_548592\|2020-0 | ···················· | ·A·················· | ···················· | ···················· | ···················· | ···················· | |
| I | 506 hCoV-19/USA/CA-CZB-4046/2020\|EPI_ISL_548593\|2020-0 | ···················· | ·A·················· | ···················· | ···················· | ···················· | ···················· | |
| I | 507 hCoV-19/USA/CA-CZB-4066/2020\|EPI_ISL_548594\|2020-0 | ···················· | ···················· | ···················· | ···················· | ···················· | ···················· | |
| I | 508 hCoV-19/USA/CA-CZB-4032/2020\|EPI_ISL_548595\|2020-0 | ···················· | ·A·················· | ···················· | ···················· | ···················· | ···················· | |
| I | 509 hCoV-19/USA/CA-CZB-4039/2020\|EPI_ISL_548596\|2020-0 | ···················· | ···················· | ···················· | ···················· | ···················· | ···················· | |

```
I    510 hCoV-19/USA/CA-CZB-4043/2020|EPI_ISL_548597|2020-0   ····················  ·A··················  ····················  ····················  ····················  ····················
I    511 hCoV-19/USA/CA-CZB-3995/2020|EPI_ISL_548598|2020-0   ····················  ····················  ····················  ····················  ····················  ····················
I    512 hCoV-19/USA/CA-CZB-4102/2020|EPI_ISL_548599|2020-0   ····················  ·A··················  ····················  ····················  ····················  ····················
I    513 hCoV-19/USA/TX-HMH-3112/2020|EPI_ISL_544000|2020-0   ····················  ····················  ····················  ····················  ····················  ····················
I    515 hCoV-19/USA/TX-HMH-3117/2020|EPI_ISL_544003|2020-0   ····················  ····················  ····················  ····················  ····················  ····················
I    516 hCoV-19/USA/TX-HMH-3118/2020|EPI_ISL_544004|2020-0   ····················  ·············A······  ····················  ····················  ····················  ····················
I    517 hCoV-19/USA/TX-HMH-3119/2020|EPI_ISL_544005|2020-0   ····················  ····················  ····················  ····················  ····················  ····················
I    518 hCoV-19/USA/TX-HMH-3120/2020|EPI_ISL_544006|2020-0   ····················  ····················  ····················  ····················  ····················  ····················
I    519 hCoV-19/USA/TX-HMH-3168/2020|EPI_ISL_544044|2020-0   ····················  ····················  ····················  ····················  ····················  ····················
I    520 hCoV-19/USA/TX-HMH-3169/2020|EPI_ISL_544045|2020-0   ····················  ·············A······  ····················  ····················  ····················  ····················
I    521 hCoV-19/USA/TX-HMH-3170/2020|EPI_ISL_544046|2020-0   ····················  ·A··················  ····················  ····················  ····················  ····················
I    522 hCoV-19/USA/TX-HMH-3171/2020|EPI_ISL_544047|2020-0   ····················  ····················  ····················  ····················  ····················  ····················
I    523 hCoV-19/USA/TX-HMH-3172/2020|EPI_ISL_544048|2020-0   ····················  ····················  ····················  ····················  ····················  ····················
I    524 hCoV-19/USA/TX-HMH-3174/2020|EPI_ISL_544049|2020-0   ····················  ····················  ····················  ····················  ····················  ····················
I    525 hCoV-19/USA/TX-HMH-3175/2020|EPI_ISL_544050|2020-0   ····················  ····················  ····················  ····················  ····················  ····················
I    526 hCoV-19/USA/TX-HMH-3176/2020|EPI_ISL_544051|2020-0   ····················  ····················  ····················  ····················  ····················  ····················
I    527 hCoV-19/USA/TX-HMH-3179/2020|EPI_ISL_544053|2020-0   ····················  ····················  ····················  ····················  ····················  ····················
I    528 hCoV-19/USA/TX-HMH-3180/2020|EPI_ISL_544054|2020-0   ····················  ····················  ····················  ····················  ····················  ····················
I    529 hCoV-19/USA/TX-HMH-3181/2020|EPI_ISL_544055|2020-0   ····················  ····················  ····················  ····················  ····················  ····················
I    530 hCoV-19/USA/TX-HMH-3182/2020|EPI_ISL_544056|2020-0   ····················  ·············A······  ····················  ····················  ····················  ····················
I    531 hCoV-19/USA/TX-HMH-3183/2020|EPI_ISL_544057|2020-0   ····················  ····················  ····················  ····················  ····················  ····················
I    532 hCoV-19/USA/TX-HMH-3185/2020|EPI_ISL_544059|2020-0   ····················  ····················  ····················  ····················  ····················  ····················
I    533 hCoV-19/USA/TX-HMH-3186/2020|EPI_ISL_544060|2020-0   ····················  ····················  ····················  ····················  ····················  ····················
I    534 hCoV-19/USA/TX-HMH-3187/2020|EPI_ISL_544061|2020-0   ····················  ····················  ····················  ····················  ····················  ····················
I    535 hCoV-19/USA/TX-HMH-3188/2020|EPI_ISL_544062|2020-0   ····················  ····················  ····················  ····················  ····················  ····················
I    536 hCoV-19/USA/TX-HMH-3189/2020|EPI_ISL_544063|2020-0   ····················  ····················  ····················  ····················  ····················  ···N·······N········
I    537 hCoV-19/USA/TX-HMH-3190/2020|EPI_ISL_544064|2020-0   ····················  ····················  ····················  ····················  ····················  ····················
I    538 hCoV-19/USA/TX-HMH-3191/2020|EPI_ISL_544065|2020-0   ····················  ····················  ····················  ····················  ····················  ····················
I    539 hCoV-19/USA/TX-HMH-3193/2020|EPI_ISL_544066|2020-0   ····················  ····················  ····················  ····················  ····················  ····················
I    540 hCoV-19/USA/TX-HMH-3194/2020|EPI_ISL_544067|2020-0   ····················  ····················  ····················  ····················  ····················  ···N················
I    541 hCoV-19/USA/TX-HMH-3195/2020|EPI_ISL_544068|2020-0   ····················  ····················  ····················  ····················  ····················  ····················
I    542 hCoV-19/USA/TX-HMH-3197/2020|EPI_ISL_544069|2020-0   ····················  ····················  ····················  ····················  ····················  ····················
I    543 hCoV-19/USA/TX-HMH-3198/2020|EPI_ISL_544070|2020-0   ····················  ····················  ····················  ····················  ····················  ····················
I    544 hCoV-19/USA/TX-HMH-3199/2020|EPI_ISL_544071|2020-0   ····················  ····················  ····················  ····················  ····················  ····················
I    545 hCoV-19/USA/TX-HMH-3203/2020|EPI_ISL_544074|2020-0   ····················  ····················  ····················  ····················  ····················  ····················
I    546 hCoV-19/USA/TX-HMH-3206/2020|EPI_ISL_544075|2020-0   ····················  ·············A······  ····················  ····················  ····················  ····················
I    548 hCoV-19/USA/TX-HMH-3209/2020|EPI_ISL_544078|2020-0   ····················  ····················  ····················  ····················  ····················  ····················
I    549 hCoV-19/USA/TX-HMH-3211/2020|EPI_ISL_544079|2020-0   ····················  ····················  ····················  ····················  ····················  ····················
I    550 hCoV-19/USA/TX-HMH-3212/2020|EPI_ISL_544080|2020-0   ····················  ····················  ····················  ····················  ····················  ····················
I    551 hCoV-19/USA/TX-HMH-3213/2020|EPI_ISL_544081|2020-0   ····················  ·············A······  ····················  ····················  ····················  ····················
I    552 hCoV-19/USA/TX-HMH-3214/2020|EPI_ISL_544082|2020-0   ····················  ····················  ····················  ····················  ····················  ····················
I    553 hCoV-19/USA/TX-HMH-3215/2020|EPI_ISL_544083|2020-0   ····················  ····················  ····················  ····················  ····················  ····················
I    554 hCoV-19/USA/TX-HMH-3216/2020|EPI_ISL_544084|2020-0   ····················  ····················  ····················  ····················  ····················  ····················
I    555 hCoV-19/USA/TX-HMH-3217/2020|EPI_ISL_544085|2020-0   ····················  ····················  ····················  ····················  ····················  ····················
I    556 hCoV-19/USA/TX-HMH-3218/2020|EPI_ISL_544086|2020-0   ····················  ····················  ····················  ····················  ····················  ····················
I    557 hCoV-19/USA/TX-HMH-3219/2020|EPI_ISL_544087|2020-0   ····················  ····················  ····················  ····················  ····················  ····················
I    558 hCoV-19/USA/TX-HMH-3220/2020|EPI_ISL_544088|2020-0   ····················  ·············A······  ····················  ····················  ····················  ····················
I    559 hCoV-19/USA/TX-HMH-3221/2020|EPI_ISL_544089|2020-0   ····················  ····················  ····················  ····················  ····················  ····················
I    560 hCoV-19/USA/TX-HMH-5594/2020|EPI_ISL_544888|2020-0   ····················  ····················  ····················  ····················  ····················  ·······N············
I    561 hCoV-19/USA/TX-HMH-5595/2020|EPI_ISL_544889|2020-0   ····················  ····················  ····················  ····················  ····················  ····················
I    562 hCoV-19/USA/TX-HMH-5598/2020|EPI_ISL_544890|2020-0   ····················  ····················  ····················  ····················  ····················  ····················
I    564 hCoV-19/USA/TX-HMH-5602/2020|EPI_ISL_544893|2020-0   ····················  ····················  ····················  ····················  ····················  ····················
I    566 hCoV-19/USA/TX-HMH-5605/2020|EPI_ISL_544896|2020-0   ····················  ····················  ····················  ····················  ····················  ····················
I    567 hCoV-19/USA/TX-HMH-5608/2020|EPI_ISL_544899|2020-0   ····················  ····················  ····················  ····················  ····················  NN··················
I    568 hCoV-19/USA/TX-HMH-5635/2020|EPI_ISL_544921|2020-0   ····················  ····················  ····················  ····················  ····················  ····················
I    569 hCoV-19/USA/TX-HMH-5636/2020|EPI_ISL_544922|2020-0   ····················  ·············A······  ····················  ····················  ····················  ····················
I    570 hCoV-19/USA/TX-HMH-5638/2020|EPI_ISL_544923|2020-0   ····················  ····················  ····················  ····················  ····················  ····················
I    571 hCoV-19/USA/TX-HMH-564/2020|EPI_ISL_544925|2020-04   ····················  ····················  ····················  ····················  ····················  ····················
I    572 hCoV-19/USA/TX-HMH-5642/2020|EPI_ISL_544928|2020-0   ····················  ····················  ····················  ····················  ····················  ····················
I    575 hCoV-19/USA/TX-HMH-5648/2020|EPI_ISL_544932|2020-0   ····················  ····················  ····················  ····················  ····················  ····················
I    576 hCoV-19/USA/TX-HMH-5649/2020|EPI_ISL_544933|2020-0   ····················  ····················  ····················  ····················  ····················  ····················
I    577 hCoV-19/USA/TX-HMH-565/2020|EPI_ISL_544934|2020-04   ····················  ····················  ····················  ····················  ····················  ····················
I    578 hCoV-19/USA/TX-HMH-5650/2020|EPI_ISL_544935|2020-0   ····················  ····················  ····················  ····················  ····················  ····················
I    579 hCoV-19/USA/TX-HMH-5651/2020|EPI_ISL_544936|2020-0   ····················  ····················  ····················  ····················  ····················  ····················
I    580 hCoV-19/USA/TX-HMH-5653/2020|EPI_ISL_544937|2020-0   ····················  ····················  ····················  ····················  ····················  ···N················
I    586 hCoV-19/USA/TX-HMH-5667/2020|EPI_ISL_544948|2020-0   ····················  ····················  ····················  ····················  ··············N·····  NN·N················   1
I    587 hCoV-19/USA/TX-HMH-567/2020|EPI_ISL_544950|2020-04   ····················  ····················  ····················  ····················  ····················  ····················
I    589 hCoV-19/USA/TX-HMH-5674/2020|EPI_ISL_544954|2020-0   ····················  ····················  ····················  ····················  ··············N·····  ····················
I    590 hCoV-19/Australia/NSW874/2020|EPI_ISL_544955|2020-   ····················  ····················  ····················  ····················  ····················  ····················
I    591 hCoV-19/Australia/NSW875/2020|EPI_ISL_544956|2020-   ····················  ····················  ····················  ····················  ····················  ····················
I    592 hCoV-19/Australia/NSW876/2020|EPI_ISL_544957|2020-   ····················  ····················  ····················  ····················  ····················  ····················
I    593 hCoV-19/Australia/NSW877/2020|EPI_ISL_544958|2020-   ····················  ····················  ····················  ····················  ····················  ····················
I    594 hCoV-19/Australia/NSW878/2020|EPI_ISL_544959|2020-   ····················  ····················  ····················  ····················  ····················  ····················
I    595 hCoV-19/Australia/NSW879/2020|EPI_ISL_544960|2020-   ····················  ····················  ····················  ····················  ····················  ····················
I    596 hCoV-19/Australia/NSW880/2020|EPI_ISL_544961|2020-   ····················  ····················  ····················  ····················  ····················  ····················
I    597 hCoV-19/Australia/NSW881/2020|EPI_ISL_544962|2020-   ····················  ····················  ····················  ····················  ····················  ····················
I    598 hCoV-19/Australia/NSW882/2020|EPI_ISL_544963|2020-   ····················  ····················  ····················  ····················  ····················  ····················
I    599 hCoV-19/Australia/NSW883/2020|EPI_ISL_544964|2020-   ····················  ····················  ····················  ····················  ····················  ····················
I    600 hCoV-19/Australia/NSW884/2020|EPI_ISL_544965|2020-   ····················  ····················  ····················  ····················  ····················  ····················
I    601 hCoV-19/Australia/NSW885/2020|EPI_ISL_544966|2020-   ····················  ····················  ····················  ····················  ····················  ····················
```

| | | | | | | | |
|---|---|---|---|---|---|---|---|
| I | 602 hCoV-19/Australia/NSW889/2020\|EPI_ISL_544970\|2020- | ···················· | ···················· | ···················· | ···················· | ···················· | ···················· |
| I | 603 hCoV-19/Australia/NSW890/2020\|EPI_ISL_544971\|2020- | ···················· | ···················· | ···················· | ···················· | ···················· | ···················· |
| I | 604 hCoV-19/Australia/NSW891/2020\|EPI_ISL_544972\|2020- | ···················· | ···················· | ···················· | ···················· | ···················· | ···················· |
| I | 605 hCoV-19/Australia/NSW892/2020\|EPI_ISL_544973\|2020- | ···················· | ···················· | ···················· | ···················· | ···················· | ···················· |
| I | 606 hCoV-19/Australia/NSW893/2020\|EPI_ISL_544974\|2020- | ···················· | ···················· | ···················· | ···················· | ···················· | ···················· |
| I | 607 hCoV-19/USA/TX-HMH-2419/2020\|EPI_ISL_545300\|2020-0 | ···················· | ·············A······ | ···················· | ···················· | ···················· | ···················· |
| I | 608 hCoV-19/USA/TX-HMH-2421/2020\|EPI_ISL_545302\|2020-0 | ···················· | ···················· | ···················· | ···················· | ···················· | ···················· |
| I | 609 hCoV-19/USA/TX-HMH-2422/2020\|EPI_ISL_545303\|2020-0 | ···················· | ···················· | ···················· | ···················· | ···················· | ···················· |
| I | 610 hCoV-19/USA/TX-HMH-2424/2020\|EPI_ISL_545305\|2020-0 | ···················· | ···················· | ···················· | ···················· | ···················· | ···················· |
| I | 611 hCoV-19/USA/TX-HMH-2425/2020\|EPI_ISL_545306\|2020-0 | ···················· | ···················· | ···················· | ···················· | ···················· | ···················· |
| I | 612 hCoV-19/USA/TX-HMH-2434/2020\|EPI_ISL_545307\|2020-0 | ···················· | ···················· | ···················· | ···················· | ···················· | ···················· |
| I | 614 hCoV-19/USA/TX-HMH-2441/2020\|EPI_ISL_545310\|2020-0 | ···················· | ···················· | ···················· | ···················· | ···················· | ···················· |
| I | 615 hCoV-19/USA/TX-HMH-2444/2020\|EPI_ISL_545311\|2020-0 | ···················· | ···················· | ···················· | ···················· | ···················· | ···················· |
| I | 616 hCoV-19/USA/TX-HMH-2445/2020\|EPI_ISL_545312\|2020-0 | ···················· | ···················· | ···················· | ···················· | ···················· | ···················· |
| I | 617 hCoV-19/USA/TX-HMH-2446/2020\|EPI_ISL_545313\|2020-0 | ···················· | ···················· | ···················· | ···················· | ···················· | ···················· |
| I | 618 hCoV-19/USA/TX-HMH-2447/2020\|EPI_ISL_545314\|2020-0 | ···················· | ···················· | ···················· | ···················· | ···················· | ···················· |
| I | 619 hCoV-19/USA/TX-HMH-2449/2020\|EPI_ISL_545315\|2020-0 | ···················· | ···················· | ···················· | ···················· | ···················· | ···················· |
| I | 620 hCoV-19/USA/TX-HMH-2450/2020\|EPI_ISL_545316\|2020-0 | ···················· | ···················· | ···················· | ···················· | ···················· | ···················· |
| I | 621 hCoV-19/USA/TX-HMH-2451/2020\|EPI_ISL_545317\|2020-0 | ···················· | ···················· | ···················· | ···················· | ···················· | ···················· |
| I | 622 hCoV-19/USA/TX-HMH-2456/2020\|EPI_ISL_545319\|2020-0 | ···················· | ···················· | ···················· | ···················· | ···················· | ···················· |
| I | 623 hCoV-19/USA/TX-HMH-2457/2020\|EPI_ISL_545320\|2020-0 | ···················· | ···················· | ···················· | ···················· | ···················· | ···················· |
| I | 624 hCoV-19/USA/TX-HMH-2459/2020\|EPI_ISL_545321\|2020-0 | ···················· | ···················· | ···················· | ···················· | ···················· | ···················· |
| I | 625 hCoV-19/USA/TX-HMH-2460/2020\|EPI_ISL_545322\|2020-0 | ···················· | ···················· | ···················· | ···················· | ···················· | ···················· |
| I | 626 hCoV-19/USA/TX-HMH-2462/2020\|EPI_ISL_545324\|2020-0 | ···················· | ···················· | ···················· | ···················· | ···················· | ···················· |
| I | 627 hCoV-19/USA/TX-HMH-2465/2020\|EPI_ISL_545325\|2020-0 | ···················· | ···················· | ···················· | ···················· | ···················· | ···················· |
| I | 628 hCoV-19/USA/TX-HMH-2466/2020\|EPI_ISL_545326\|2020-0 | ···················· | ···················· | ···················· | ···················· | ···················· | ···················· |
| I | 629 hCoV-19/USA/TX-HMH-2470/2020\|EPI_ISL_545327\|2020-0 | ···················· | ···················· | ···················· | ···················· | ···················· | ···················· |
| I | 630 hCoV-19/USA/TX-HMH-2471/2020\|EPI_ISL_545328\|2020-0 | ···················· | ···················· | ···················· | ···················· | ···················· | ···················· |
| I | 631 hCoV-19/USA/TX-HMH-2472/2020\|EPI_ISL_545329\|2020-0 | ···················· | ···················· | ···················· | ···················· | ···················· | ···················· |
| I | 632 hCoV-19/USA/TX-HMH-2473/2020\|EPI_ISL_545330\|2020-0 | ···················· | ···················· | ···················· | ···················· | ···················· | ···················· |
| I | 633 hCoV-19/USA/TX-HMH-2474/2020\|EPI_ISL_545331\|2020-0 | ···················· | ···················· | ···················· | ···················· | ···················· | ···················· |
| I | 634 hCoV-19/USA/TX-HMH-2475/2020\|EPI_ISL_545332\|2020-0 | ···················· | ···················· | ···················· | ···················· | ···················· | ···················· |
| I | 635 hCoV-19/USA/TX-HMH-2478/2020\|EPI_ISL_545333\|2020-0 | ···················· | ···················· | ···················· | ···················· | ···················· | ···················· |
| I | 636 hCoV-19/USA/TX-HMH-2482/2020\|EPI_ISL_545334\|2020-0 | ···················· | ···················· | ···················· | ···················· | ···················· | ···················· |
| I | 637 hCoV-19/USA/TX-HMH-2483/2020\|EPI_ISL_545335\|2020-0 | ···················· | ···················· | ···················· | ···················· | ···················· | ···················· |
| I | 638 hCoV-19/USA/TX-HMH-2486/2020\|EPI_ISL_545336\|2020-0 | ···················· | ···················· | ···················· | ···················· | ···················· | ···················· |
| I | 639 hCoV-19/USA/TX-HMH-2489/2020\|EPI_ISL_545337\|2020-0 | ···················· | ···················· | ···················· | ···················· | ···················· | ···················· |
| I | 640 hCoV-19/USA/TX-HMH-2545/2020\|EPI_ISL_545375\|2020-0 | ···················· | ···················· | ···················· | ···················· | ···················· | ···················· |
| I | 641 hCoV-19/USA/TX-HMH-2546/2020\|EPI_ISL_545376\|2020-0 | ···················· | ···················· | ···················· | ···················· | ···················· | ···················· |
| I | 642 hCoV-19/USA/TX-HMH-2547/2020\|EPI_ISL_545377\|2020-0 | ···················· | ···················· | ···················· | ···················· | ···················· | ···················· |
| I | 643 hCoV-19/USA/TX-HMH-2549/2020\|EPI_ISL_545378\|2020-0 | ···················· | ···················· | ···················· | ···················· | ···················· | ·······N············ |
| I | 644 hCoV-19/USA/TX-HMH-2553/2020\|EPI_ISL_545381\|2020-0 | ···················· | ···················· | ···················· | ···················· | ···················· | ···················· |
| I | 645 hCoV-19/USA/TX-HMH-2556/2020\|EPI_ISL_545383\|2020-0 | ···················· | ···················· | ···················· | ···················· | ···················· | ···················· |
| I | 646 hCoV-19/USA/TX-HMH-2557/2020\|EPI_ISL_545384\|2020-0 | ···················· | ···················· | ···················· | ···················· | ···················· | ···················· |
| I | 647 hCoV-19/USA/TX-HMH-2558/2020\|EPI_ISL_545385\|2020-0 | ···················· | ···················· | ···················· | ···················· | ···················· | ···················· |
| I | 648 hCoV-19/USA/TX-HMH-2561/2020\|EPI_ISL_545387\|2020-0 | ···················· | ···················· | ···················· | ···················· | ···················· | ···················· |
| I | 649 hCoV-19/USA/TX-HMH-2562/2020\|EPI_ISL_545388\|2020-0 | ···················· | ···················· | ···················· | ···················· | ···················· | ···················· |
| I | 650 hCoV-19/USA/TX-HMH-2564/2020\|EPI_ISL_545389\|2020-0 | ···················· | ···················· | ···················· | ···················· | ···················· | ···················· |
| I | 651 hCoV-19/USA/TX-HMH-2565/2020\|EPI_ISL_545390\|2020-0 | ···················· | ···················· | ···················· | ···················· | ···················· | ···················· |
| I | 652 hCoV-19/USA/TX-HMH-2566/2020\|EPI_ISL_545391\|2020-0 | ···················· | ···················· | ···················· | ···················· | ···················· | ···················· |
| I | 653 hCoV-19/USA/TX-HMH-2567/2020\|EPI_ISL_545392\|2020-0 | ···················· | ···················· | ···················· | ···················· | ···················· | ···················· |
| I | 654 hCoV-19/USA/TX-HMH-2568/2020\|EPI_ISL_545393\|2020-0 | ···················· | ···················· | ···················· | ···················· | ···················· | ···················· |
| I | 655 hCoV-19/USA/TX-HMH-2570/2020\|EPI_ISL_545394\|2020-0 | ···················· | ···················· | ···················· | ···················· | ···················· | ···················· |
| I | 656 hCoV-19/USA/TX-HMH-2572/2020\|EPI_ISL_545395\|2020-0 | ···················· | ···················· | ···················· | ···················· | ···················· | ···················· |
| I | 657 hCoV-19/USA/TX-HMH-2573/2020\|EPI_ISL_545396\|2020-0 | ···················· | ···················· | ···················· | ···················· | ···················· | ···················· |
| I | 658 hCoV-19/USA/TX-HMH-2574/2020\|EPI_ISL_545397\|2020-0 | ···················· | ···················· | ···················· | ···················· | ···················· | ···················· |
| I | 659 hCoV-19/USA/TX-HMH-2575/2020\|EPI_ISL_545398\|2020-0 | ···················· | ···················· | ···················· | ···················· | ···················· | ···················· |
| I | 660 hCoV-19/USA/TX-HMH-2576/2020\|EPI_ISL_545399\|2020-0 | ···················· | ···················· | ···················· | ···················· | ···················· | ···················· |
| I | 663 hCoV-19/USA/TX-HMH-7214/2020\|EPI_ISL_546603\|2020-0 | ···················· | ···················· | ···················· | ···················· | ···················· | ···················· |
| I | 664 hCoV-19/USA/TX-HMH-7215/2020\|EPI_ISL_546604\|2020-0 | ···················· | ···················· | ···················· | ···················· | ···················· | ···················· |
| I | 667 hCoV-19/USA/TX-HMH-7218/2020\|EPI_ISL_546607\|2020-0 | ···················· | ···················· | ···················· | ···················· | ···················· | ···················· |
| I | 668 hCoV-19/USA/TX-HMH-7222/2020\|EPI_ISL_546610\|2020-0 | ···················· | ···················· | ···················· | ···················· | ···················· | ···················· |
| I | 670 hCoV-19/USA/TX-HMH-7227/2020\|EPI_ISL_546612\|2020-0 | ···················· | ···················· | ···················· | ···················· | ···················· | ···················· |
| I | 671 hCoV-19/USA/TX-HMH-7228/2020\|EPI_ISL_546613\|2020-0 | ···················· | ···················· | ···················· | ···················· | ···················· | ···················· |
| I | 672 hCoV-19/USA/TX-HMH-7230/2020\|EPI_ISL_546614\|2020-0 | ···················· | ···················· | ···················· | ···················· | ···················· | NN·················· |
| I | 673 hCoV-19/USA/TX-HMH-7233/2020\|EPI_ISL_546615\|2020-0 | ···················· | ···················· | ···················· | ···················· | ···················· | ···················· |
| I | 674 hCoV-19/USA/TX-HMH-7235/2020\|EPI_ISL_546617\|2020-0 | ···················· | ···················· | ···················· | ···················· | ···················· | ···················· |
| I | 676 hCoV-19/USA/TX-HMH-7238/2020\|EPI_ISL_546619\|2020-0 | ···················· | ···················· | ···················· | ···················· | ···················· | ···················· |
| I | 677 hCoV-19/USA/TX-HMH-7240/2020\|EPI_ISL_546621\|2020-0 | ···················· | ···················· | ···················· | ···················· | ···················· | ···················· |
| I | 678 hCoV-19/USA/TX-HMH-7244/2020\|EPI_ISL_546623\|2020-0 | ···················· | ···················· | ···················· | ···················· | ···················· | ···················· |
| I | 679 hCoV-19/USA/TX-HMH-7246/2020\|EPI_ISL_546624\|2020-0 | ···················· | ···················· | ···················· | ···················· | ···················· | ···················· |
| I | 683 hCoV-19/USA/TX-HMH-7259/2020\|EPI_ISL_546631\|2020-0 | ···················· | ···················· | ···················· | ···················· | ···················· | ···················· |
| I | 684 hCoV-19/USA/TX-HMH-7261/2020\|EPI_ISL_546632\|2020-0 | ···················· | ···················· | ···················· | ···················· | ···················· | ···················· |
| I | 685 hCoV-19/USA/TX-HMH-7266/2020\|EPI_ISL_546636\|2020-0 | ···················· | ···················· | ···················· | ···················· | ···················· | ···················· |
| I | 687 hCoV-19/USA/TX-HMH-7270/2020\|EPI_ISL_546639\|2020-0 | ···················· | ···················· | ···················· | ···················· | ···················· | ···················· |
| I | 692 hCoV-19/USA/TX-HMH-7283/2020\|EPI_ISL_546647\|2020-0 | ···················· | ···················· | ···················· | ···················· | ··············N····· | ···················· |
| I | 693 hCoV-19/USA/TX-HMH-7292/2020\|EPI_ISL_546650\|2020-0 | ···················· | ···················· | ···················· | ···················· | ···················· | ···················· |
| I | 695 hCoV-19/USA/TX-HMH-7295/2020\|EPI_ISL_546652\|2020-0 | ···················· | ···················· | ···················· | ···················· | ···················· | ···················· |
| I | 696 hCoV-19/USA/TX-HMH-7296/2020\|EPI_ISL_546653\|2020-0 | ···················· | ···················· | ···················· | ···················· | ···················· | ···················· |
| I | 697 hCoV-19/USA/TX-HMH-730/2020\|EPI_ISL_546656\|2020-04 | ···················· | ···················· | ···················· | ···················· | ···················· | ···················· |

```
I    698 hCoV-19/USA/TX-HMH-7300/2020|EPI_ISL_546657|2020-0    ····················  ····················  ····················  ····················  ····················  ····················
I    700 hCoV-19/USA/TX-HMH-7302/2020|EPI_ISL_546659|2020-0    ····················  ····················  ····················  ····················  ····················  ····················
I    701 hCoV-19/USA/TX-HMH-7303/2020|EPI_ISL_546660|2020-0    ····················  ·A··················  ····················  ····················  ····················  ····················
I    704 hCoV-19/USA/TX-HMH-7310/2020|EPI_ISL_546667|2020-0    ····················  ····················  ····················  ····················  ····················  ····················
I    706 hCoV-19/USA/TX-HMH-5907/2020|EPI_ISL_547100|2020-0    ····················  ····················  ····················  ····················  ····················  ····················
I    709 hCoV-19/USA/TX-HMH-5912/2020|EPI_ISL_547104|2020-0    ····················  ······A·············  ····················  ····················  ····················  ····················
I    711 hCoV-19/USA/TX-HMH-5916/2020|EPI_ISL_547106|2020-0    ····················  ····················  ····················  ····················  ····················  ····················
I    712 hCoV-19/USA/TX-HMH-5917/2020|EPI_ISL_547107|2020-0    ····················  ····················  ····················  ····················  ····················  ····················
I    713 hCoV-19/USA/TX-HMH-592/2020|EPI_ISL_547109|2020-04    ····················  ····················  ····················  ····················  ····················  ····················
I    715 hCoV-19/USA/TX-HMH-5944/2020|EPI_ISL_547111|2020-0    ····················  ····················  ····················  ····················  ····················  ····················
I    718 hCoV-19/USA/TX-HMH-5953/2020|EPI_ISL_547118|2020-0    ····················  ····················  ····················  ····················  ····················  ····················
I    719 hCoV-19/USA/TX-HMH-5955/2020|EPI_ISL_547119|2020-0    ····················  ····················  ····················  ····················  ····················  ····················
I    720 hCoV-19/Czech Republic/NRL_8118-3/2020|EPI_ISL_547    ····················  ····················  ····················  ····················  ····················  ····················
I    721 hCoV-19/Czech Republic/NRL_8118-4/2020|EPI_ISL_547    ····················  ····················  ····················  ····················  ····················  ····················
I    722 hCoV-19/Czech Republic/NRL_8118-5/2020|EPI_ISL_547    ····················  ····················  ····················  ····················  ····················  ····················
I    723 hCoV-19/New Zealand/20CV0054/2020|EPI_ISL_547969|2    ····················  ····················  ····················  ····················  ····················  ····················
I    724 hCoV-19/New Zealand/20CV0055/2020|EPI_ISL_547970|2    ····················  ····················  ····················  ····················  ····················  ····················
I    725 hCoV-19/New Zealand/20CV0057/2020|EPI_ISL_547971|2    ····················  ····················  ····················  ····················  ····················  ····················
I    726 hCoV-19/New Zealand/20CV0058/2020|EPI_ISL_547972|2    ····················  ····················  ····················  ····················  ····················  ····················
I    727 hCoV-19/New Zealand/20CV0059/2020|EPI_ISL_547973|2    ····················  ····················  ····················  ····················  ····················  ····················
I    728 hCoV-19/New Zealand/20CV0060/2020|EPI_ISL_547974|2    ····················  ····················  ····················  ····················  ····················  ····················
I    729 hCoV-19/New Zealand/20CV0061/2020|EPI_ISL_547975|2    ····················  ····················  ····················  ····················  ····················  ····················
I    730 hCoV-19/New Zealand/20CV0067/2020|EPI_ISL_547976|2    ····················  ····················  ····················  ····················  ····················  ····················
I    731 hCoV-19/New Zealand/20CV0068/2020|EPI_ISL_547977|2    ····················  ····················  ····················  ····················  ····················  ····················
I    732 hCoV-19/New Zealand/20CV0069/2020|EPI_ISL_547978|2    ····················  ····················  ····················  ····················  ····················  ····················
I    733 hCoV-19/New Zealand/20CV0070/2020|EPI_ISL_547979|2    ····················  ····················  ····················  ····················  ····················  ····················
I    734 hCoV-19/New Zealand/20CV0075/2020|EPI_ISL_547981|2    ····················  ····················  ····················  ····················  ····················  ····················
I    735 hCoV-19/New Zealand/20CV0076/2020|EPI_ISL_547982|2    ····················  ····················  ····················  ····················  ····················  ····················
I    736 hCoV-19/New Zealand/20CV0077/2020|EPI_ISL_547983|2    ····················  ····················  ····················  ····················  ····················  ····················
I    737 hCoV-19/New Zealand/20CV0078/2020|EPI_ISL_547984|2    ····················  ····················  ····················  ····················  ····················  ····················
I    738 hCoV-19/New Zealand/20CV0079/2020|EPI_ISL_547985|2    ····················  ····················  ····················  ····················  ····················  ····················
I    739 hCoV-19/New Zealand/20CV0093/2020|EPI_ISL_547993|2    ····················  ····················  ····················  ····················  ····················  ····················
I    740 hCoV-19/New Zealand/20CV0094/2020|EPI_ISL_547994|2    ····················  ····················  ····················  ····················  ····················  ····················
I    741 hCoV-19/New Zealand/20CV0095/2020|EPI_ISL_547995|2    ····················  ····················  ····················  ····················  ····················  ····················
I    742 hCoV-19/New Zealand/20CV0102/2020|EPI_ISL_547996|2    ····················  ····················  ····················  ····················  ····················  ····················
I    743 hCoV-19/New Zealand/20CV0103/2020|EPI_ISL_547997|2    ····················  ····················  ····················  ····················  ····················  ····················
I    744 hCoV-19/New Zealand/20CV0105/2020|EPI_ISL_547998|2    ····················  ····················  ····················  ····················  ····················  ····················
I    745 hCoV-19/New Zealand/20CV0106/2020|EPI_ISL_547999|2    ····················  ····················  ····················  ····················  ····················  ····················
I    746 hCoV-19/USA/CA-CZB-3768/2020|EPI_ISL_548402|2020-0    ····················  ·A··················  ····················  ····················  ····················  ····················
I    747 hCoV-19/USA/CA-CZB-3738/2020|EPI_ISL_548404|2020-0    ····················  ····················  ····················  ····················  ····················  ····················
I    748 hCoV-19/USA/CA-CZB-3778/2020|EPI_ISL_548406|2020-0    ····················  ····················  ····················  ····················  ····················  ····················
I    749 hCoV-19/USA/CA-CZB-3731/2020|EPI_ISL_548408|2020-0    ····················  ·A··················  ····················  ····················  ····················  ····················
I    750 hCoV-19/USA/CA-CZB-3800/2020|EPI_ISL_548409|2020-0    ····················  ····················  ····················  ····················  ····················  ····················
I    751 hCoV-19/USA/CA-CZB-3783/2020|EPI_ISL_548410|2020-0    ····················  ····················  ····················  ····················  ····················  ····················
I    760 hCoV-19/USA/CA-CZB-3801/2020|EPI_ISL_548430|2020-0    ····················  ·A··················  ····················  ····················  ····················  ····················
I    764 hCoV-19/USA/CA-CZB-3724/2020|EPI_ISL_548439|2020-0    ····················  ·A··················  ····················  ····················  ····················  ····················
I    765 hCoV-19/USA/CA-CZB-3770/2020|EPI_ISL_548442|2020-0    ····················  ····················  ····················  ····················  ····················  ····················
I    766 hCoV-19/USA/CA-CZB-3773/2020|EPI_ISL_548443|2020-0    ····················  ····················  ····················  ····················  ····················  ····················
I    768 hCoV-19/USA/CA-CZB-3679/2020|EPI_ISL_548447|2020-0    ····················  ·A··················  ····················  ····················  ····················  ····················
I    769 hCoV-19/USA/TX-HMH-3223/2020|EPI_ISL_544090|2020-0    ····················  ····················  ····················  ····················  ····················  ····················
I    770 hCoV-19/USA/TX-HMH-3224/2020|EPI_ISL_544091|2020-0    ····················  ·············A······  ····················  ····················  ····················  ····················
I    771 hCoV-19/USA/TX-HMH-3225/2020|EPI_ISL_544092|2020-0    ····················  ····················  ····················  ····················  ····················  ······NN············
I    772 hCoV-19/USA/TX-HMH-3226/2020|EPI_ISL_544093|2020-0    ····················  ·············A······  ····················  ····················  ····················  ····················
I    773 hCoV-19/USA/TX-HMH-3227/2020|EPI_ISL_544094|2020-0    ····················  ····················  ····················  ····················  ····················  ····················
I    774 hCoV-19/USA/TX-HMH-3228/2020|EPI_ISL_544095|2020-0    ····················  ····················  ····················  ····················  ····················  ····················
I    775 hCoV-19/USA/TX-HMH-3229/2020|EPI_ISL_544096|2020-0    ····················  ····················  ····················  ····················  ····················  ····················
I    776 hCoV-19/USA/TX-HMH-3279/2020|EPI_ISL_544097|2020-0    ····················  ····················  ····················  ····················  ····················  ····················
I    777 hCoV-19/USA/TX-HMH-3280/2020|EPI_ISL_544098|2020-0    ····················  ····················  ····················  ····················  ····················  ····················
I    780 hCoV-19/USA/TX-HMH-5612/2020|EPI_ISL_544903|2020-0    ····················  ····················  ····················  ····················  ····················  ····················
I    781 hCoV-19/USA/TX-HMH-5615/2020|EPI_ISL_544904|2020-0    ····················  ·············A······  ····················  ····················  ····················  ····················
I    782 hCoV-19/USA/TX-HMH-5616/2020|EPI_ISL_544905|2020-0    ····················  ····················  ····················  ····················  ····················  ····················
I    783 hCoV-19/USA/TX-HMH-5618/2020|EPI_ISL_544906|2020-0    ····················  ·············A······  ····················  ····················  ····················  ····················
I    784 hCoV-19/USA/TX-HMH-5619/2020|EPI_ISL_544907|2020-0    ····················  ····················  ····················  ····················  ····················  ····················
I    785 hCoV-19/USA/TX-HMH-5623/2020|EPI_ISL_544910|2020-0    ····················  ····················  ····················  ····················  ····················  ····················
I    787 hCoV-19/USA/TX-HMH-5627/2020|EPI_ISL_544913|2020-0    ····················  ····················  ····················  ····················  ····················  ······NN············
I    788 hCoV-19/USA/TX-HMH-5629/2020|EPI_ISL_544915|2020-0    ····················  ····················  ····················  ····················  ····················  ····················
I    789 hCoV-19/USA/TX-HMH-563/2020|EPI_ISL_544916|2020-04    ····················  ····················  ····················  ····················  ····················  ····················
I    790 hCoV-19/USA/TX-HMH-5630/2020|EPI_ISL_544917|2020-0    ····················  ····················  ····················  ····················  ····················  ····················
I    791 hCoV-19/Australia/NSW886/2020|EPI_ISL_544967|2020-    ····················  ····················  ····················  ····················  ····················  ····················
I    792 hCoV-19/Australia/NSW887/2020|EPI_ISL_544968|2020-    ····················  ····················  ····················  ····················  ····················  ····················
I    793 hCoV-19/Australia/NSW888/2020|EPI_ISL_544969|2020-    ····················  ····················  ····················  ····················  ····················  ····················
I    794 hCoV-19/Australia/NSW894/2020|EPI_ISL_544975|2020-    ····················  ····················  ····················  ····················  ····················  ····················
I    795 hCoV-19/Australia/NSW895/2020|EPI_ISL_544976|2020-    ····················  ····················  ····················  ····················  ····················  ····················
I    796 hCoV-19/Australia/NSW896/2020|EPI_ISL_544977|2020-    ····················  ····················  ····················  ····················  ····················  ····················
I    797 hCoV-19/Australia/NSW897/2020|EPI_ISL_544978|2020-    ····················  ····················  ····················  ····················  ····················  ····················
I    798 hCoV-19/Australia/NSW898/2020|EPI_ISL_544979|2020-    ····················  ····················  ····················  ····················  ····················  ····················
I    799 hCoV-19/Australia/NSW899/2020|EPI_ISL_544980|2020-    ····················  ····················  ····················  ····················  ····················  ····················
I    800 hCoV-19/Australia/NSW900/2020|EPI_ISL_544981|2020-    ····················  ····················  ····················  ····················  ····················  ····················
I    801 hCoV-19/Australia/NSW901/2020|EPI_ISL_544982|2020-    ····················  ····················  ····················  ····················  ····················  ····················
I    802 hCoV-19/Australia/NSW902/2020|EPI_ISL_544983|2020-    ····················  ····················  ····················  ····················  ····················  ····················
```

| | | | | | | | | |
|---|---|---|---|---|---|---|---|---|
| I | 803 | hCoV-19/Australia/NSW903/2020\|EPI_ISL_544984\|2020- | ···················· | ···················· | ···················· | ···················· | ···················· | ···················· |
| I | 804 | hCoV-19/Australia/NSW904/2020\|EPI_ISL_544985\|2020- | ···················· | ···················· | ···················· | ···················· | ···················· | ···················· |
| I | 805 | hCoV-19/Australia/NSW905/2020\|EPI_ISL_544986\|2020- | ···················· | ···················· | ···················· | ···················· | ···················· | ···················· |
| I | 806 | hCoV-19/Australia/NSW906/2020\|EPI_ISL_544987\|2020- | ···················· | ···················· | ···················· | ···················· | ···················· | ···················· |
| I | 807 | hCoV-19/Australia/NSW907/2020\|EPI_ISL_544988\|2020- | ···················· | ···················· | ···················· | ···················· | ···················· | ···················· |
| I | 808 | hCoV-19/Australia/NSW908/2020\|EPI_ISL_544989\|2020- | ········T··········· | ···················· | ···················· | ···················· | ···················· | ···················· |
| I | 809 | hCoV-19/Australia/NSW909/2020\|EPI_ISL_544990\|2020- | ···················· | ···················· | ···················· | ···················· | ···················· | ···················· |
| I | 810 | hCoV-19/Australia/NSW910/2020\|EPI_ISL_544991\|2020- | ···················· | ···················· | ···················· | ···················· | ···················· | ···················· |
| I | 811 | hCoV-19/Australia/NSW911/2020\|EPI_ISL_544992\|2020- | ···················· | ···················· | ···················· | ···················· | ···················· | ···················· |
| I | 812 | hCoV-19/Australia/NSW912/2020\|EPI_ISL_544993\|2020- | ···················· | ···················· | ···················· | ···················· | ···················· | ···················· |
| I | 813 | hCoV-19/Australia/NSW913/2020\|EPI_ISL_544994\|2020- | ···················· | ···················· | ···················· | ···················· | ···················· | ···················· |
| I | 814 | hCoV-19/Australia/NSW914/2020\|EPI_ISL_544995\|2020- | ···················· | ···················· | ···················· | ···················· | ···················· | ···················· |
| I | 815 | hCoV-19/Australia/NSW915/2020\|EPI_ISL_544996\|2020- | ···················· | ···················· | ···················· | ···················· | ···················· | ···················· |
| I | 816 | hCoV-19/Australia/NSW916/2020\|EPI_ISL_544997\|2020- | ···················· | ···················· | ···················· | ···················· | ···················· | ···················· |
| I | 817 | hCoV-19/Australia/NSW917/2020\|EPI_ISL_544998\|2020- | ···················· | ···················· | ···················· | ···················· | ···················· | ···················· |
| I | 818 | hCoV-19/Australia/NSW918/2020\|EPI_ISL_544999\|2020- | ···················· | ···················· | ···················· | ···················· | ···················· | ···················· |
| I | 819 | hCoV-19/USA/TX-HMH-2268/2020\|EPI_ISL_545217\|2020-0 | ···················· | ···················· | ···················· | ···················· | ···················· | ···················· |
| I | 820 | hCoV-19/USA/TX-HMH-2276/2020\|EPI_ISL_545218\|2020-0 | ···················· | ···················· | ···················· | ···················· | ···················· | ···················· |
| I | 821 | hCoV-19/USA/TX-HMH-2277/2020\|EPI_ISL_545219\|2020-0 | ···················· | ···················· | ···················· | ···················· | ···················· | ···················· |
| I | 822 | hCoV-19/USA/TX-HMH-2278/2020\|EPI_ISL_545220\|2020-0 | ···················· | ···················· | ···················· | ···················· | ···················· | ···················· |
| I | 823 | hCoV-19/USA/TX-HMH-2279/2020\|EPI_ISL_545221\|2020-0 | ···················· | ···················· | ···················· | ···················· | ···················· | ···················· |
| I | 824 | hCoV-19/USA/TX-HMH-2282/2020\|EPI_ISL_545223\|2020-0 | ···················· | ···················· | ···················· | ···················· | ···················· | ···················· |
| I | 825 | hCoV-19/USA/TX-HMH-2285/2020\|EPI_ISL_545224\|2020-0 | ···················· | ···················· | ···················· | ···················· | ···················· | ···A················ |
| I | 826 | hCoV-19/USA/TX-HMH-2287/2020\|EPI_ISL_545225\|2020-0 | ···················· | ···················· | ···················· | ···················· | ···················· | ···················· |
| I | 827 | hCoV-19/USA/TX-HMH-2288/2020\|EPI_ISL_545226\|2020-0 | ···················· | ······A············· | ···················· | ···················· | ···················· | ···················· |
| I | 828 | hCoV-19/USA/TX-HMH-2290/2020\|EPI_ISL_545227\|2020-0 | ···················· | ···················· | ···················· | ···················· | ···················· | ···················· |
| I | 829 | hCoV-19/USA/TX-HMH-2291/2020\|EPI_ISL_545228\|2020-0 | ···················· | ···················· | ···················· | ···················· | ···················· | ···················· |
| I | 830 | hCoV-19/USA/TX-HMH-2293/2020\|EPI_ISL_545229\|2020-0 | ···················· | ···················· | ···················· | ···················· | ···················· | ···················· |
| I | 831 | hCoV-19/USA/TX-HMH-2295/2020\|EPI_ISL_545230\|2020-0 | ···················· | ···················· | ···················· | ···················· | ···················· | ···················· |
| I | 832 | hCoV-19/USA/TX-HMH-2296/2020\|EPI_ISL_545231\|2020-0 | ···················· | ···················· | ···················· | ···················· | ···················· | ···················· |
| I | 833 | hCoV-19/USA/TX-HMH-2300/2020\|EPI_ISL_545233\|2020-0 | ···················· | ···················· | ···················· | ···················· | ···················· | ···················· |
| I | 834 | hCoV-19/USA/TX-HMH-2301/2020\|EPI_ISL_545234\|2020-0 | ···················· | ···················· | ···················· | ···················· | ···················· | ···················· |
| I | 835 | hCoV-19/USA/TX-HMH-2305/2020\|EPI_ISL_545235\|2020-0 | ···················· | ···················· | ···················· | ···················· | ···················· | ···················· |
| I | 836 | hCoV-19/USA/TX-HMH-2306/2020\|EPI_ISL_545236\|2020-0 | ···················· | ···················· | ···················· | ···················· | ···················· | ···················· |
| I | 837 | hCoV-19/USA/TX-HMH-2307/2020\|EPI_ISL_545237\|2020-0 | ···················· | ···················· | ···················· | ···················· | ···················· | ···················· |
| I | 838 | hCoV-19/USA/TX-HMH-2308/2020\|EPI_ISL_545238\|2020-0 | ···················· | ···················· | ···················· | ···················· | ···················· | ···················· |
| I | 839 | hCoV-19/USA/TX-HMH-2310/2020\|EPI_ISL_545239\|2020-0 | ···················· | ···················· | ···················· | ···················· | ···················· | NN·················· |
| I | 840 | hCoV-19/USA/TX-HMH-2318/2020\|EPI_ISL_545240\|2020-0 | ···················· | ···················· | ···················· | ···················· | ···················· | ···················· |
| I | 841 | hCoV-19/USA/TX-HMH-2319/2020\|EPI_ISL_545241\|2020-0 | ···················· | ···················· | ···················· | ···················· | ···················· | ···················· |
| I | 842 | hCoV-19/USA/TX-HMH-2320/2020\|EPI_ISL_545242\|2020-0 | ···················· | ···················· | ···················· | ···················· | ···················· | ···················· |
| I | 843 | hCoV-19/USA/TX-HMH-2323/2020\|EPI_ISL_545243\|2020-0 | ···················· | ···················· | ···················· | ···················· | ···················· | ···················· |
| I | 844 | hCoV-19/USA/TX-HMH-2325/2020\|EPI_ISL_545244\|2020-0 | ···················· | ···················· | ···················· | ···················· | ···················· | ···················· |
| I | 845 | hCoV-19/USA/TX-HMH-2327/2020\|EPI_ISL_545246\|2020-0 | ···················· | ···················· | ···················· | ···················· | ···················· | ···················· |
| I | 846 | hCoV-19/USA/TX-HMH-2328/2020\|EPI_ISL_545247\|2020-0 | ···················· | ···················· | ···················· | ···················· | ···················· | ···················· |
| I | 847 | hCoV-19/USA/TX-HMH-2330/2020\|EPI_ISL_545249\|2020-0 | ···················· | ···················· | ···················· | ···················· | ···················· | ···················· |
| I | 848 | hCoV-19/USA/TX-HMH-2333/2020\|EPI_ISL_545251\|2020-0 | ···················· | ···················· | ···················· | ···················· | ···················· | ···················· |
| I | 849 | hCoV-19/USA/TX-HMH-2342/2020\|EPI_ISL_545255\|2020-0 | ···················· | ···················· | ···················· | ···················· | ···················· | ···················· |
| I | 850 | hCoV-19/USA/TX-HMH-2346/2020\|EPI_ISL_545256\|2020-0 | ···················· | ···················· | ···················· | ···················· | ···················· | ···················· |
| I | 851 | hCoV-19/USA/TX-HMH-2350/2020\|EPI_ISL_545258\|2020-0 | ···················· | ···················· | ···················· | ···················· | ···················· | ···················· |
| I | 852 | hCoV-19/USA/TX-HMH-2351/2020\|EPI_ISL_545259\|2020-0 | ···················· | ···················· | ···················· | ···················· | ···················· | ···················· |
| I | 853 | hCoV-19/USA/TX-HMH-2352/2020\|EPI_ISL_545260\|2020-0 | ···················· | ···················· | ···················· | ···················· | ···················· | ···················· |
| I | 854 | hCoV-19/USA/TX-HMH-2356/2020\|EPI_ISL_545261\|2020-0 | ···················· | ···················· | ···················· | ···················· | ···················· | ·······N············ |
| I | 855 | hCoV-19/USA/TX-HMH-2357/2020\|EPI_ISL_545262\|2020-0 | ···················· | ···················· | ···················· | ···················· | ···················· | ···················· |
| I | 856 | hCoV-19/USA/TX-HMH-2359/2020\|EPI_ISL_545263\|2020-0 | ···················· | ···················· | ···················· | ···················· | ···················· | ···················· |
| I | 857 | hCoV-19/USA/TX-HMH-2361/2020\|EPI_ISL_545264\|2020-0 | ···················· | ···················· | ···················· | ···················· | ···················· | ···················· |
| I | 858 | hCoV-19/USA/TX-HMH-2366/2020\|EPI_ISL_545266\|2020-0 | ···················· | ···················· | ···················· | ···················· | ···················· | ···················· |
| I | 859 | hCoV-19/USA/TX-HMH-2368/2020\|EPI_ISL_545267\|2020-0 | ···················· | ···················· | ···················· | ···················· | ···················· | ···················· |
| I | 860 | hCoV-19/USA/TX-HMH-2371/2020\|EPI_ISL_545269\|2020-0 | ···················· | ···················· | ···················· | ···················· | ···················· | ···················· |
| I | 861 | hCoV-19/USA/TX-HMH-2374/2020\|EPI_ISL_545270\|2020-0 | ···················· | ··············C····· | ···················· | ···················· | ···················· | ···················· |
| I | 862 | hCoV-19/USA/TX-HMH-2375/2020\|EPI_ISL_545271\|2020-0 | ···················· | ······A············· | ···················· | ···················· | ···················· | ···················· |
| I | 863 | hCoV-19/USA/TX-HMH-2377/2020\|EPI_ISL_545272\|2020-0 | ···················· | ···················· | ···················· | ···················· | ···················· | ···················· |
| I | 864 | hCoV-19/USA/TX-HMH-2379/2020\|EPI_ISL_545273\|2020-0 | ···················· | ···················· | ···················· | ···················· | ···················· | ···················· |
| I | 865 | hCoV-19/USA/TX-HMH-2381/2020\|EPI_ISL_545274\|2020-0 | ···················· | ······A············· | ···················· | ···················· | ···················· | ···················· |
| I | 866 | hCoV-19/USA/TX-HMH-2383/2020\|EPI_ISL_545276\|2020-0 | ···················· | ···················· | ···················· | ···················· | ···················· | ···················· |
| I | 867 | hCoV-19/USA/TX-HMH-2384/2020\|EPI_ISL_545277\|2020-0 | ···················· | ···················· | ···················· | ···················· | ···················· | ···················· |
| I | 868 | hCoV-19/USA/TX-HMH-2386/2020\|EPI_ISL_545279\|2020-0 | ···················· | ···················· | ···················· | ···················· | ···················· | ···················· |
| I | 869 | hCoV-19/USA/TX-HMH-2389/2020\|EPI_ISL_545281\|2020-0 | ···················· | ···················· | ···················· | ···················· | ···················· | ···················· |
| I | 870 | hCoV-19/USA/TX-HMH-2390/2020\|EPI_ISL_545282\|2020-0 | ···················· | ···················· | ···················· | ···················· | ···················· | ···················· |
| I | 871 | hCoV-19/USA/TX-HMH-2392/2020\|EPI_ISL_545283\|2020-0 | ···················· | ······A············· | ···················· | ···················· | ···················· | ···················· |
| I | 872 | hCoV-19/USA/TX-HMH-2395/2020\|EPI_ISL_545284\|2020-0 | ···················· | ···················· | ···················· | ···················· | ···················· | ·······N············ |
| I | 873 | hCoV-19/USA/TX-HMH-2397/2020\|EPI_ISL_545285\|2020-0 | ···················· | ···················· | ···················· | ···················· | ···················· | ···················· |
| I | 874 | hCoV-19/USA/TX-HMH-2401/2020\|EPI_ISL_545288\|2020-0 | ···················· | ···················· | ···················· | ···················· | ···················· | ···················· |
| I | 875 | hCoV-19/USA/TX-HMH-2402/2020\|EPI_ISL_545289\|2020-0 | ···················· | ···················· | ···················· | ···················· | ···················· | ···················· |
| I | 881 | hCoV-19/USA/TX-HMH-716/2020\|EPI_ISL_546555\|2020-04 | ···················· | ···················· | ···················· | ···················· | ···················· | ···················· |
| I | 882 | hCoV-19/USA/TX-HMH-7160/2020\|EPI_ISL_546556\|2020-0 | ···················· | ···················· | ···················· | ···················· | ···················· | ···················· |
| I | 883 | hCoV-19/USA/TX-HMH-7161/2020\|EPI_ISL_546557\|2020-0 | ···················· | ·············A······ | ···················· | ···················· | ···················· | ···················· |
| I | 885 | hCoV-19/USA/TX-HMH-7168/2020\|EPI_ISL_546563\|2020-0 | ···················· | ···················· | ···················· | ···················· | ···················· | ···················· |
| I | 886 | hCoV-19/USA/TX-HMH-7169/2020\|EPI_ISL_546564\|2020-0 | ···················· | ···················· | ···················· | ···················· | ···················· | ···················· |
| I | 887 | hCoV-19/USA/TX-HMH-717/2020\|EPI_ISL_546565\|2020-04 | ···················· | ···················· | ···················· | ···················· | ···················· | ···················· |
| I | 888 | hCoV-19/USA/TX-HMH-7170/2020\|EPI_ISL_546566\|2020-0 | ···················· | ···················· | ···················· | ···················· | ···················· | ···················· |

| | | | | | | | | |
|---|---|---|---|---|---|---|---|---|
| I | 889 hCoV-19/USA/TX-HMH-7171/2020\|EPI_ISL_546567\|2020-0 | ···················· | ·············A······ | ···················· | ···················· | ···················· | ···················· | |
| I | 891 hCoV-19/USA/TX-HMH-7173/2020\|EPI_ISL_546569\|2020-0 | ···················· | ···················· | ···················· | ···················· | ···················· | ···················· | |
| I | 892 hCoV-19/USA/TX-HMH-7174/2020\|EPI_ISL_546570\|2020-0 | ···················· | ···················· | ···················· | ···················· | ···················· | ···················· | |
| I | 893 hCoV-19/USA/TX-HMH-7175/2020\|EPI_ISL_546571\|2020-0 | ···················· | ···················· | ···················· | ···················· | ···················· | ···················· | |
| I | 895 hCoV-19/USA/TX-HMH-7179/2020\|EPI_ISL_546574\|2020-0 | ···················· | ·············A······ | ···················· | ···················· | ···················· | ···················· | |
| I | 896 hCoV-19/USA/TX-HMH-718/2020\|EPI_ISL_546575\|2020-04 | ···················· | ···················· | ···················· | ···················· | ···················· | ···················· | |
| I | 898 hCoV-19/USA/TX-HMH-7185/2020\|EPI_ISL_546581\|2020-0 | ···················· | ···················· | ···················· | ···················· | ······T············· | ···N················ | |
| I | 901 hCoV-19/USA/TX-HMH-7191/2020\|EPI_ISL_546585\|2020-0 | ···················· | ···················· | ···················· | ···················· | ···················· | ···················· | |
| I | 902 hCoV-19/USA/TX-HMH-7192/2020\|EPI_ISL_546586\|2020-0 | ···················· | ···················· | ···················· | ···················· | ···················· | ···················· | |
| I | 905 hCoV-19/USA/TX-HMH-720/2020\|EPI_ISL_546593\|2020-04 | ···················· | ···················· | ···················· | ···················· | ···················· | ···················· | |
| I | 906 hCoV-19/USA/TX-HMH-7203/2020\|EPI_ISL_546594\|2020-0 | ···················· | ···················· | ···················· | ···················· | ···················· | ···················· | |
| I | 907 hCoV-19/USA/TX-HMH-7207/2020\|EPI_ISL_546598\|2020-0 | ···················· | ·············A······ | ···················· | ···················· | ···················· | ···················· | |
| I | 908 hCoV-19/USA/TX-HMH-7208/2020\|EPI_ISL_546599\|2020-0 | ···················· | ···················· | ···················· | ···················· | ···················· | ···················· | |
| I | 909 hCoV-19/USA/TX-HMH-7355/2020\|EPI_ISL_546700\|2020-0 | ···················· | ···················· | ···················· | ···················· | ···················· | ···················· | |
| I | 912 hCoV-19/USA/TX-HMH-5958/2020\|EPI_ISL_547122\|2020-0 | ···················· | ···················· | ···················· | ···················· | ···················· | ···················· | |
| I | 913 hCoV-19/USA/TX-HMH-5959/2020\|EPI_ISL_547123\|2020-0 | ···················· | ·A·················· | ···················· | ···················· | ···················· | ···················· | |
| I | 914 hCoV-19/USA/TX-HMH-5960/2020\|EPI_ISL_547124\|2020-0 | ···················· | ···················· | ···················· | ···················· | ···················· | ···················· | |
| I | 916 hCoV-19/USA/TX-HMH-5962/2020\|EPI_ISL_547126\|2020-0 | ···················· | ···················· | ···················· | ···················· | ···················· | ···················· | |
| I | 921 hCoV-19/USA/TX-HMH-597/2020\|EPI_ISL_547132\|2020-04 | ···················· | ···················· | ···················· | ···················· | ···················· | ···················· | |
| I | 923 hCoV-19/USA/TX-HMH-5972/2020\|EPI_ISL_547135\|2020-0 | ···················· | ···················· | ···················· | ···················· | ··············N····· | ·······N············ | |
| I | 925 hCoV-19/USA/TX-HMH-5978/2020\|EPI_ISL_547139\|2020-0 | ···················· | ···················· | ···················· | ···················· | ···················· | ···················· | |
| I | 926 hCoV-19/USA/TX-HMH-598/2020\|EPI_ISL_547140\|2020-04 | ···················· | ···················· | ···················· | ···················· | ···················· | ···················· | |
| I | 928 hCoV-19/USA/TX-HMH-5984/2020\|EPI_ISL_547144\|2020-0 | ···················· | ···················· | ···················· | ···················· | ···················· | NN·N················ | 1 |
| I | 929 hCoV-19/USA/TX-HMH-5986/2020\|EPI_ISL_547146\|2020-0 | ···················· | ···················· | ···················· | ···················· | ··············N····· | ············N······· | |
| I | 932 hCoV-19/USA/TX-HMH-5993/2020\|EPI_ISL_547152\|2020-0 | ···················· | ···················· | ···················· | ···················· | ···················· | ···················· | |
| I | 933 hCoV-19/USA/TX-HMH-5998/2020\|EPI_ISL_547155\|2020-0 | ···················· | ···················· | ···················· | ···················· | ···················· | ···················· | |
| I | 934 hCoV-19/USA/TX-HMH-600/2020\|EPI_ISL_547156\|2020-04 | ···················· | ···················· | ···················· | ···················· | ···················· | ···················· | |
| I | 935 hCoV-19/USA/TX-HMH-6001/2020\|EPI_ISL_547158\|2020-0 | ···················· | ·············A······ | ···················· | ···················· | ···················· | ···················· | |
| I | 936 hCoV-19/USA/TX-HMH-6003/2020\|EPI_ISL_547159\|2020-0 | ···················· | ·············A······ | ···················· | ···················· | ···················· | ···················· | |
| I | 937 hCoV-19/USA/TX-HMH-6004/2020\|EPI_ISL_547160\|2020-0 | ···················· | ···················· | ···················· | ···················· | ···················· | ···················· | |
| I | 939 hCoV-19/USA/TX-HMH-6007/2020\|EPI_ISL_547162\|2020-0 | ···················· | ···················· | ···················· | ···················· | ···················· | ···················· | |
| I | 940 hCoV-19/USA/TX-HMH-6008/2020\|EPI_ISL_547163\|2020-0 | ···················· | ···················· | ···················· | ···················· | ···················· | ···················· | |
| I | 941 hCoV-19/USA/TX-HMH-601/2020\|EPI_ISL_547165\|2020-04 | ···················· | ···················· | ···················· | ···················· | ···················· | ···················· | |
| I | 942 hCoV-19/USA/TX-HMH-6010/2020\|EPI_ISL_547166\|2020-0 | ···················· | ···················· | ···················· | ···················· | ···················· | ···················· | |
| I | 943 hCoV-19/USA/TX-HMH-6013/2020\|EPI_ISL_547168\|2020-0 | ···················· | ···················· | ···················· | ···················· | ···················· | ···················· | |
| I | 944 hCoV-19/USA/TX-HMH-6014/2020\|EPI_ISL_547169\|2020-0 | ···················· | ······A············· | ···················· | ···················· | ···················· | ···················· | |
| I | 945 hCoV-19/USA/TX-HMH-6016/2020\|EPI_ISL_547170\|2020-0 | ···················· | ···················· | ···················· | ···················· | ···················· | ···················· | |
| I | 947 hCoV-19/USA/TX-HMH-602/2020\|EPI_ISL_547172\|2020-04 | ···················· | ···················· | ···················· | ···················· | ···················· | ···················· | |
| I | 948 hCoV-19/USA/TX-HMH-6020/2020\|EPI_ISL_547173\|2020-0 | ···················· | ·············A······ | ···················· | ···················· | ···················· | ···················· | |
| I | 950 hCoV-19/USA/TX-HMH-6025/2020\|EPI_ISL_547175\|2020-0 | ···················· | ···················· | ···················· | ···················· | ···················· | ···················· | |
| I | 951 hCoV-19/USA/TX-HMH-6026/2020\|EPI_ISL_547176\|2020-0 | ···················· | ···················· | ···················· | ···················· | ···················· | ···················· | |
| I | 952 hCoV-19/USA/TX-HMH-6028/2020\|EPI_ISL_547177\|2020-0 | ···················· | ···················· | ···················· | ···················· | ···················· | ···················· | |
| I | 953 hCoV-19/USA/TX-HMH-6029/2020\|EPI_ISL_547178\|2020-0 | ···················· | ···················· | ···················· | ···················· | ···················· | ···················· | |
| I | 954 hCoV-19/USA/TX-HMH-6034/2020\|EPI_ISL_547180\|2020-0 | ···················· | ·············A······ | ···················· | ···················· | ···················· | ···················· | |
| I | 955 hCoV-19/USA/TX-HMH-6035/2020\|EPI_ISL_547181\|2020-0 | ···················· | ···················· | ···················· | ···················· | ···················· | ···················· | |
| I | 956 hCoV-19/USA/TX-HMH-6036/2020\|EPI_ISL_547182\|2020-0 | ···················· | ···················· | ···················· | ···················· | ···················· | ···················· | |
| I | 957 hCoV-19/USA/TX-HMH-6037/2020\|EPI_ISL_547183\|2020-0 | ···················· | ···················· | ···················· | ···················· | ···················· | ···················· | |
| I | 958 hCoV-19/USA/TX-HMH-6038/2020\|EPI_ISL_547184\|2020-0 | ···················· | ···················· | ···················· | ···················· | ···················· | ···················· | |
| I | 962 hCoV-19/USA/TX-HMH-6043/2020\|EPI_ISL_547188\|2020-0 | ···················· | ···················· | ···················· | ···················· | ···················· | ···················· | |
| I | 963 hCoV-19/USA/TX-HMH-6046/2020\|EPI_ISL_547190\|2020-0 | ···················· | ······A············· | ···················· | ···················· | ···················· | NN·················· | |
| I | 964 hCoV-19/USA/TX-HMH-6047/2020\|EPI_ISL_547191\|2020-0 | ···················· | ···················· | ···················· | ···················· | ···················· | ···················· | |
| I | 966 hCoV-19/USA/TX-HMH-6052/2020\|EPI_ISL_547195\|2020-0 | ···················· | ···················· | ···················· | ···················· | ···················· | ···················· | |
| I | 968 hCoV-19/USA/CA-CZB-3338/2020\|EPI_ISL_548300\|2020-0 | ···················· | ···················· | ···················· | ···················· | ···················· | ···················· | |
| I | 969 hCoV-19/USA/CA-CZB-3361/2020\|EPI_ISL_548301\|2020-0 | ···················· | ···················· | ···················· | ···················· | ···················· | ···················· | |
| I | 970 hCoV-19/USA/CA-CZB-3292/2020\|EPI_ISL_548302\|2020-0 | ···················· | ···················· | ···················· | ···················· | ···················· | ···················· | |
| I | 971 hCoV-19/USA/CA-CZB-3293/2020\|EPI_ISL_548303\|2020-0 | ···················· | ·A·················· | ···················· | ···················· | ···················· | ···················· | |
| I | 972 hCoV-19/USA/CA-CZB-3322/2020\|EPI_ISL_548304\|2020-0 | ···················· | ·A·················· | ···················· | ···················· | ···················· | ···················· | |
| I | 973 hCoV-19/USA/CA-CZB-3334/2020\|EPI_ISL_548305\|2020-0 | ···················· | ·A·················· | ···················· | ···················· | ···················· | ···················· | |
| I | 974 hCoV-19/USA/CA-CZB-3341/2020\|EPI_ISL_548306\|2020-0 | ···················· | ·A·················· | ···················· | ···················· | ···················· | ···················· | |
| I | 975 hCoV-19/USA/CA-CZB-3287/2020\|EPI_ISL_548307\|2020-0 | ···················· | ·A·················· | ···················· | ···················· | ···················· | ···················· | |
| I | 976 hCoV-19/USA/CA-CZB-3355/2020\|EPI_ISL_548308\|2020-0 | ···················· | ···················· | ···················· | ···················· | ···················· | ···················· | |
| I | 977 hCoV-19/USA/CA-CZB-3353/2020\|EPI_ISL_548309\|2020-0 | ···················· | ···················· | ···················· | ···················· | ···················· | ···················· | |
| I | 978 hCoV-19/USA/CA-CZB-3285/2020\|EPI_ISL_548310\|2020-0 | ···················· | ·A·················· | ···················· | ···················· | ···················· | ···················· | |
| I | 979 hCoV-19/USA/CA-CZB-3299/2020\|EPI_ISL_548311\|2020-0 | ···················· | ···················· | ···········T········ | ···················· | ···················· | ···················· | |
| I | 980 hCoV-19/USA/CA-CZB-3356/2020\|EPI_ISL_548312\|2020-0 | ···················· | ···················· | ···················· | ···················· | ···················· | ···················· | |
| I | 981 hCoV-19/USA/CA-CZB-3330/2020\|EPI_ISL_548313\|2020-0 | ···················· | ···················· | ···················· | ···················· | ···················· | ··············W····· | |
| I | 982 hCoV-19/USA/CA-CZB-3271/2020\|EPI_ISL_548314\|2020-0 | ···················· | ···················· | ···················· | ···················· | ···················· | ···················· | |
| I | 983 hCoV-19/USA/CA-CZB-3354/2020\|EPI_ISL_548315\|2020-0 | ···················· | ···················· | ···················· | ···················· | ···················· | ···················· | |
| I | 984 hCoV-19/USA/CA-CZB-3314/2020\|EPI_ISL_548316\|2020-0 | ···················· | ···················· | ···················· | ···················· | ···················· | ···················· | |
| I | 985 hCoV-19/USA/CA-CZB-3335/2020\|EPI_ISL_548317\|2020-0 | ···················· | ···················· | ···················· | ···················· | ···················· | ···················· | |
| I | 986 hCoV-19/USA/CA-CZB-3784/2020\|EPI_ISL_548450\|2020-0 | ···················· | ···················· | ···················· | ···················· | ···················· | ···················· | |
| I | 987 hCoV-19/USA/CA-CZB-3676/2020\|EPI_ISL_548452\|2020-0 | ···················· | ···················· | ···················· | ···················· | ···················· | ···················· | |
| I | 988 hCoV-19/USA/CA-CZB-3720/2020\|EPI_ISL_548453\|2020-0 | ···················· | ···················· | ···················· | ···················· | NNN················· | ···················· | 1 |
| I | 989 hCoV-19/USA/CA-CZB-3766/2020\|EPI_ISL_548455\|2020-0 | ···················· | ·A·················· | ···················· | ···················· | ···················· | ···················· | |
| I | 990 hCoV-19/USA/CA-CZB-3723/2020\|EPI_ISL_548457\|2020-0 | ···················· | ···················· | ···················· | ···················· | ···················· | ···················· | |
| I | 991 hCoV-19/USA/CA-CZB-3775/2020\|EPI_ISL_548459\|2020-0 | ···················· | ···················· | ···················· | ···················· | ···················· | ···················· | |
| I | 992 hCoV-19/USA/CA-CZB-3737/2020\|EPI_ISL_548460\|2020-0 | ···················· | ·A·················· | ···················· | ···················· | ···················· | ···················· | |
| I | 993 hCoV-19/USA/CA-CZB-3769/2020\|EPI_ISL_548461\|2020-0 | ···················· | ···················· | ···················· | ···················· | ···················· | ···················· | |
| I | 994 hCoV-19/USA/CA-CZB-3745/2020\|EPI_ISL_548463\|2020-0 | ···················· | ·A·················· | ···················· | ···················· | ···················· | ···················· | |
| I | 995 hCoV-19/USA/CA-CZB-3733/2020\|EPI_ISL_548464\|2020-0 | ···················· | ···················· | ···················· | ···················· | ···················· | ···················· | |

| | | | | | | | | |
|---|---|---|---|---|---|---|---|---|
| I | 996 | hCoV-19/USA/CA-CZB-3719/2020\|EPI_ISL_548470\|2020-0 | ···················· | ·A·················· | ···················· | ···················· | ··N················· | ···················· |
| I | 997 | hCoV-19/USA/CA-CZB-3781/2020\|EPI_ISL_548471\|2020-0 | ···················· | ···················· | ···················· | ···················· | ···················· | ···················· |
| I | 998 | hCoV-19/USA/CA-CZB-3763/2020\|EPI_ISL_548473\|2020-0 | ···················· | ···················· | ···················· | ···················· | ···················· | ···················· |
| I | 999 | hCoV-19/USA/CA-CZB-3683/2020\|EPI_ISL_548474\|2020-0 | ···················· | ···················· | ···················· | ···················· | ···················· | ···················· |
| I | 1000 | hCoV-19/USA/CA-CZB-3762/2020\|EPI_ISL_548475\|2020-0 | ···················· | ···················· | ···················· | ···················· | ···················· | ···················· |
| I | 1001 | hCoV-19/USA/CA-CZB-3776/2020\|EPI_ISL_548476\|2020-0 | ···················· | ···················· | ···················· | ···················· | ···················· | ···················· |
| I | 1002 | hCoV-19/USA/CA-CZB-3678/2020\|EPI_ISL_548477\|2020-0 | ···················· | ···················· | ···················· | ···················· | ···················· | ···················· |
| I | 1003 | hCoV-19/USA/CA-CZB-3698/2020\|EPI_ISL_548478\|2020-0 | ···················· | ·A·················· | ···················· | ···················· | ···················· | ···················· |
| I | 1004 | hCoV-19/USA/CA-CZB-3677/2020\|EPI_ISL_548479\|2020-0 | ···················· | ···················· | ···················· | ···················· | ···················· | ···················· |
| I | 1005 | hCoV-19/USA/CA-CZB-3764/2020\|EPI_ISL_548480\|2020-0 | ···················· | ···················· | ···················· | ···················· | ···················· | ···················· |
| I | 1006 | hCoV-19/USA/CA-CZB-3787/2020\|EPI_ISL_548481\|2020-0 | ···················· | ···················· | ···················· | ···················· | ···················· | ···················· |
| I | 1008 | hCoV-19/USA/CA-CZB-3793/2020\|EPI_ISL_548483\|2020-0 | ···················· | ·A·················· | ···················· | ···················· | ···················· | ···················· |
| I | 1009 | hCoV-19/USA/CA-CZB-3759/2020\|EPI_ISL_548484\|2020-0 | ···················· | ···················· | ···················· | ···················· | ···················· | ···················· |
| I | 1010 | hCoV-19/USA/CA-CZB-3746/2020\|EPI_ISL_548485\|2020-0 | ···················· | ···················· | ···················· | ···················· | ···················· | ···················· |
| I | 1011 | hCoV-19/USA/CA-CZB-3681/2020\|EPI_ISL_548486\|2020-0 | ···················· | ···················· | ···················· | ···················· | ···················· | ···················· |
| I | 1012 | hCoV-19/USA/CA-CZB-3779/2020\|EPI_ISL_548487\|2020-0 | ···················· | ···················· | ···················· | ···················· | ···················· | ···················· |
| I | 1013 | hCoV-19/USA/CA-CZB-3741/2020\|EPI_ISL_548488\|2020-0 | ···················· | ···················· | ···················· | ···················· | ···················· | ···················· |
| I | 1014 | hCoV-19/USA/CA-CZB-3792/2020\|EPI_ISL_548489\|2020-0 | ···················· | ···················· | ···················· | ···················· | ···················· | ···················· |
| I | 1015 | hCoV-19/USA/CA-CZB-3689/2020\|EPI_ISL_548490\|2020-0 | ···················· | ···················· | ···················· | ···················· | ···················· | ···················· |
| I | 1016 | hCoV-19/USA/CA-CZB-3928/2020\|EPI_ISL_548491\|2020-0 | ···················· | ···················· | ···················· | ···················· | ···················· | ···················· |
| I | 1017 | hCoV-19/USA/CA-CZB-3946/2020\|EPI_ISL_548492\|2020-0 | ···················· | ···················· | ···················· | ···················· | ···················· | ···················· |
| I | 1018 | hCoV-19/USA/CA-CZB-3898/2020\|EPI_ISL_548493\|2020-0 | ···················· | ·A·················· | ···················· | ···················· | ···················· | ···················· |
| I | 1019 | hCoV-19/USA/CA-CZB-3882/2020\|EPI_ISL_548494\|2020-0 | ···················· | ···················· | ···················· | ···················· | ···················· | ···················· |
| I | 1020 | hCoV-19/USA/CA-CZB-3926/2020\|EPI_ISL_548495\|2020-0 | ···················· | ···················· | ···················· | ···················· | ···················· | ···················· |
| I | 1021 | hCoV-19/USA/CA-CZB-3889/2020\|EPI_ISL_548496\|2020-0 | ···················· | ···················· | ···················· | ···················· | ···················· | ···················· |
| I | 1022 | hCoV-19/USA/CA-CZB-3866/2020\|EPI_ISL_548497\|2020-0 | ···················· | ·A·················· | ···················· | ···················· | ···················· | ···················· |
| I | 1023 | hCoV-19/USA/CA-CZB-3891/2020\|EPI_ISL_548498\|2020-0 | ···················· | ···················· | ···················· | ···················· | ···················· | ···················· |
| I | 1024 | hCoV-19/USA/CA-CZB-3919/2020\|EPI_ISL_548499\|2020-0 | ···················· | ···················· | ···················· | ···················· | ···················· | ···················· |
| I | 1025 | hCoV-19/USA/TX-HMH-5424/2020\|EPI_ISL_544766\|2020-0 | ···················· | ···················· | ···················· | ···················· | ···················· | ···················· |
| I | 1026 | hCoV-19/USA/TX-HMH-5425/2020\|EPI_ISL_544767\|2020-0 | ···················· | ···················· | ···················· | ···················· | ···················· | ···················· |
| I | 1027 | hCoV-19/USA/TX-HMH-5428/2020\|EPI_ISL_544768\|2020-0 | ···················· | ···················· | ···················· | ···················· | ···················· | ···················· |
| I | 1029 | hCoV-19/USA/TX-HMH-5434/2020\|EPI_ISL_544772\|2020-0 | ···················· | ···················· | ···················· | ···················· | ···················· | ···················· |
| I | 1030 | hCoV-19/USA/TX-HMH-5435/2020\|EPI_ISL_544773\|2020-0 | ···················· | ···················· | ···················· | ···················· | ···················· | ···················· |
| I | 1031 | hCoV-19/USA/TX-HMH-5437/2020\|EPI_ISL_544775\|2020-0 | ···················· | ···················· | ···················· | ···················· | ···················· | ···················· |
| I | 1037 | hCoV-19/USA/TX-HMH-5444/2020\|EPI_ISL_544781\|2020-0 | ···················· | ···················· | ···················· | ···················· | ···················· | ···················· |
| I | 1040 | hCoV-19/USA/TX-HMH-5449/2020\|EPI_ISL_544784\|2020-0 | ···················· | ·············A······ | ···················· | ···················· | ···················· | ···················· |
| I | 1044 | hCoV-19/USA/TX-HMH-5468/2020\|EPI_ISL_544795\|2020-0 | ···················· | ···················· | ···················· | ···················· | ···················· | ···················· |
| I | 1045 | hCoV-19/USA/TX-HMH-547/2020\|EPI_ISL_544796\|2020-04 | ···················· | ···················· | ···················· | ···················· | ···················· | ···················· |
| I | 1046 | hCoV-19/USA/TX-HMH-5471/2020\|EPI_ISL_544797\|2020-0 | ···················· | ···················· | ···················· | ···················· | ···················· | ···················· |
| I | 1047 | hCoV-19/USA/TX-HMH-5472/2020\|EPI_ISL_544798\|2020-0 | ···················· | ··········A········· | ···················· | ···················· | ···················· | ···················· |
| I | 1048 | hCoV-19/USA/TX-HMH-5473/2020\|EPI_ISL_544799\|2020-0 | ···················· | ·············A······ | ···················· | ···················· | ···················· | ···················· |
| I | 1049 | hCoV-19/USA/TX-HMH-5474/2020\|EPI_ISL_544800\|2020-0 | ···················· | ······A············· | ···················· | ···················· | ···················· | ···················· |
| I | 1052 | hCoV-19/USA/TX-HMH-548/2020\|EPI_ISL_544805\|2020-04 | ···················· | ···················· | ···················· | ···················· | ···················· | ···················· |
| I | 1053 | hCoV-19/USA/TX-HMH-5480/2020\|EPI_ISL_544806\|2020-0 | ···················· | ···················· | ···················· | ···················· | ···················· | ···················· |
| I | 1054 | hCoV-19/USA/TX-HMH-5481/2020\|EPI_ISL_544807\|2020-0 | ···················· | ···················· | ···················· | ···················· | ···················· | ···················· |
| I | 1055 | hCoV-19/USA/TX-HMH-549/2020\|EPI_ISL_544812\|2020-04 | ···················· | ···················· | ···················· | ···················· | ···················· | ···················· |
| I | 1057 | hCoV-19/USA/TX-HMH-5492/2020\|EPI_ISL_544815\|2020-0 | ···················· | ···················· | ···················· | ···················· | ···················· | ···················· |
| I | 1060 | hCoV-19/USA/TX-HMH-5497/2020\|EPI_ISL_544820\|2020-0 | ···················· | ···················· | ···················· | ···················· | ···················· | ···················· |
| I | 1062 | hCoV-19/USA/TX-HMH-5499/2020\|EPI_ISL_544822\|2020-0 | ···················· | ·············A······ | ···················· | ···················· | ···················· | ···················· |
| I | 1068 | hCoV-19/USA/TX-HMH-5507/2020\|EPI_ISL_544830\|2020-0 | ···················· | ···················· | ···················· | ···················· | ···················· | ···················· |
| I | 1073 | hCoV-19/USA/TX-HMH-5511/2020\|EPI_ISL_544835\|2020-0 | ···················· | ···················· | ···················· | ···················· | ···················· | ···················· |
| I | 1074 | hCoV-19/USA/TX-HMH-5516/2020\|EPI_ISL_544838\|2020-0 | ···················· | ···················· | ···················· | ···················· | ···················· | ···················· |
| I | 1075 | hCoV-19/USA/TX-HMH-5517/2020\|EPI_ISL_544839\|2020-0 | ···················· | ···················· | ···················· | ···················· | ···················· | ···················· |
| I | 1076 | hCoV-19/USA/TX-HMH-5518/2020\|EPI_ISL_544840\|2020-0 | ···················· | ···················· | ···················· | ···················· | ···················· | ···················· |
| I | 1077 | hCoV-19/USA/TX-HMH-5520/2020\|EPI_ISL_544841\|2020-0 | ···················· | ···················· | ···················· | ···················· | ···················· | ···················· |
| I | 1083 | hCoV-19/USA/TX-HMH-5538/2020\|EPI_ISL_544848\|2020-0 | ···················· | ···················· | ···················· | ···················· | ···················· | ···················· |
| I | 1085 | hCoV-19/USA/TX-HMH-5541/2020\|EPI_ISL_544850\|2020-0 | ···················· | ···················· | ···················· | ···················· | ···················· | ···················· |
| I | 1087 | hCoV-19/USA/TX-HMH-5545/2020\|EPI_ISL_544852\|2020-0 | ···················· | ···················· | ···················· | ···················· | ···················· | ···················· |
| I | 1088 | hCoV-19/USA/TX-HMH-5546/2020\|EPI_ISL_544853\|2020-0 | ···················· | ···················· | ···················· | ···················· | ···················· | ···················· |
| I | 1093 | hCoV-19/USA/TX-HMH-5558/2020\|EPI_ISL_544860\|2020-0 | ···················· | ···················· | ···················· | ···················· | ··············N····· | ··············N····· |
| I | 1096 | hCoV-19/USA/TX-HMH-5566/2020\|EPI_ISL_544867\|2020-0 | ···················· | ···················· | ···················· | ···················· | ···················· | ···················· |
| I | 1097 | hCoV-19/USA/TX-HMH-557/2020\|EPI_ISL_544868\|2020-04 | ···················· | ···················· | ···················· | ···················· | ···················· | ···················· |
| I | 1101 | hCoV-19/USA/TX-HMH-5575/2020\|EPI_ISL_544873\|2020-0 | ···················· | ·············A······ | ···················· | ···················· | ···················· | ···················· |
| I | 1103 | hCoV-19/USA/TX-HMH-5578/2020\|EPI_ISL_544875\|2020-0 | ···················· | ···················· | ···················· | ···················· | ···················· | ···················· |
| I | 1104 | hCoV-19/USA/TX-HMH-5579/2020\|EPI_ISL_544876\|2020-0 | ···················· | ···················· | ···················· | ···················· | ···················· | ···················· |
| I | 1106 | hCoV-19/USA/TX-HMH-5583/2020\|EPI_ISL_544879\|2020-0 | ···················· | ···················· | ···················· | ···················· | ···················· | ···················· |
| I | 1107 | hCoV-19/USA/TX-HMH-5586/2020\|EPI_ISL_544882\|2020-0 | ···················· | ···················· | ···················· | ···················· | ···················· | ···················· |
| I | 1108 | hCoV-19/USA/TX-HMH-5587/2020\|EPI_ISL_544883\|2020-0 | ···················· | ···················· | ···················· | ···················· | ···················· | ···················· |
| I | 1109 | hCoV-19/USA/TX-HMH-5588/2020\|EPI_ISL_544884\|2020-0 | ···················· | ···················· | ···················· | ···················· | ···················· | ···················· |
| I | 1111 | hCoV-19/USA/TX-HMH-5592/2020\|EPI_ISL_544886\|2020-0 | ···················· | ···················· | ···················· | ···················· | ···················· | ···················· |
| I | 1113 | hCoV-19/USA/TX-HMH-2246/2020\|EPI_ISL_545201\|2020-0 | ···················· | ···················· | ···················· | ···················· | ···················· | ···················· |
| I | 1114 | hCoV-19/USA/TX-HMH-2250/2020\|EPI_ISL_545204\|2020-0 | ···················· | ······A············· | ···················· | ···················· | ···················· | ···················· |
| I | 1115 | hCoV-19/USA/TX-HMH-2252/2020\|EPI_ISL_545205\|2020-0 | ···················· | ···················· | ···················· | ···················· | ···················· | ···················· |
| I | 1116 | hCoV-19/USA/TX-HMH-2253/2020\|EPI_ISL_545206\|2020-0 | ···················· | ···················· | ···················· | ···················· | ···················· | ···················· |
| I | 1117 | hCoV-19/USA/TX-HMH-2254/2020\|EPI_ISL_545207\|2020-0 | ···················· | ···················· | ···················· | ···················· | ···················· | ···················· |
| I | 1118 | hCoV-19/USA/TX-HMH-2255/2020\|EPI_ISL_545208\|2020-0 | ···················· | ···················· | ···················· | ···················· | ···················· | ···················· |
| I | 1119 | hCoV-19/USA/TX-HMH-2260/2020\|EPI_ISL_545210\|2020-0 | ···················· | ···················· | ···················· | ···················· | ···················· | ···················· |
| I | 1120 | hCoV-19/USA/TX-HMH-2265/2020\|EPI_ISL_545214\|2020-0 | ···················· | ···················· | ···················· | ···················· | ···················· | ···················· |
| I | 1121 | hCoV-19/USA/TX-HMH-2267/2020\|EPI_ISL_545216\|2020-0 | ···················· | ···················· | ···················· | ···················· | ···················· | ···················· |
| I | 1122 | hCoV-19/USA/TX-HMH-2403/2020\|EPI_ISL_545290\|2020-0 | ···················· | ···················· | ···················· | ···················· | ···················· | ···················· |

| | | | | | | | | |
|---|---|---|---|---|---|---|---|---|
| I | 1123 hCoV-19/USA/TX-HMH-2404/2020\|EPI_ISL_545291\|2020-0 | ···················· | ······A············· | ···················· | ···················· | ···················· | ···················· | |
| I | 1124 hCoV-19/USA/TX-HMH-2412/2020\|EPI_ISL_545295\|2020-0 | ···················· | ···················· | ···················· | ···················· | ···················· | ···················· | |
| I | 1125 hCoV-19/USA/TX-HMH-2413/2020\|EPI_ISL_545296\|2020-0 | ···················· | ···················· | ···················· | ···················· | ···················· | ···················· | |
| I | 1126 hCoV-19/USA/TX-HMH-2415/2020\|EPI_ISL_545297\|2020-0 | ···················· | ···················· | ···················· | ···················· | ···················· | ···················· | |
| I | 1127 hCoV-19/USA/TX-HMH-2416/2020\|EPI_ISL_545298\|2020-0 | ···················· | ···················· | ···················· | ···················· | ···················· | ···················· | |
| I | 1128 hCoV-19/USA/TX-HMH-2418/2020\|EPI_ISL_545299\|2020-0 | ···················· | ···················· | ···················· | ···················· | ···················· | ···················· | |
| I | 1129 hCoV-19/USA/TX-HMH-7096/2020\|EPI_ISL_546500\|2020-0 | ···················· | ···················· | ···················· | ···················· | ···················· | ···················· | |
| I | 1130 hCoV-19/USA/TX-HMH-7099/2020\|EPI_ISL_546502\|2020-0 | ···················· | ···················· | ···················· | ···················· | ···················· | ···················· | |
| I | 1131 hCoV-19/USA/TX-HMH-7100/2020\|EPI_ISL_546503\|2020-0 | ···················· | ···················· | ···················· | ···················· | ···················· | ···················· | |
| I | 1132 hCoV-19/USA/TX-HMH-7101/2020\|EPI_ISL_546504\|2020-0 | ···················· | ···················· | ···················· | ···················· | ···················· | ···················· | |
| I | 1133 hCoV-19/USA/TX-HMH-7103/2020\|EPI_ISL_546505\|2020-0 | ···················· | ···················· | ···················· | ···················· | ···················· | ···················· | |
| I | 1134 hCoV-19/USA/TX-HMH-7104/2020\|EPI_ISL_546506\|2020-0 | ···················· | ···················· | ···················· | ···················· | ···················· | ···················· | |
| I | 1135 hCoV-19/USA/TX-HMH-7105/2020\|EPI_ISL_546507\|2020-0 | ···················· | ·············A······ | ···················· | ···················· | ···················· | ···················· | |
| I | 1136 hCoV-19/USA/TX-HMH-7107/2020\|EPI_ISL_546509\|2020-0 | ···················· | ···················· | ···················· | ···················· | ···················· | ···················· | |
| I | 1137 hCoV-19/USA/TX-HMH-7108/2020\|EPI_ISL_546510\|2020-0 | ···················· | ···················· | ···················· | ···················· | ···················· | ···················· | |
| I | 1138 hCoV-19/USA/TX-HMH-711/2020\|EPI_ISL_546511\|2020-04 | ···················· | ···················· | ···················· | ···················· | ···················· | ···················· | |
| I | 1139 hCoV-19/USA/TX-HMH-7110/2020\|EPI_ISL_546512\|2020-0 | ···················· | ···················· | ···················· | ···················· | ···················· | ···················· | |
| I | 1140 hCoV-19/USA/TX-HMH-7114/2020\|EPI_ISL_546515\|2020-0 | ···················· | ···················· | ···················· | ···················· | ···················· | ···················· | |
| I | 1141 hCoV-19/USA/TX-HMH-7116/2020\|EPI_ISL_546517\|2020-0 | ···················· | ···················· | ···················· | ···················· | ···················· | ···················· | |
| I | 1142 hCoV-19/USA/TX-HMH-7117/2020\|EPI_ISL_546518\|2020-0 | ···················· | ···················· | ···················· | ···················· | ···················· | ···················· | |
| I | 1143 hCoV-19/USA/TX-HMH-7118/2020\|EPI_ISL_546519\|2020-0 | ···················· | ···················· | ···················· | ···················· | ···················· | ···················· | |
| I | 1144 hCoV-19/USA/TX-HMH-7119/2020\|EPI_ISL_546520\|2020-0 | ···················· | ···················· | ···················· | ···················· | ·········N····N····· | ···N··NN············ | 1 |
| I | 1146 hCoV-19/USA/TX-HMH-7121/2020\|EPI_ISL_546522\|2020-0 | ···················· | ·············A······ | ···················· | ···················· | ···················· | ···················· | |
| I | 1148 hCoV-19/USA/TX-HMH-7123/2020\|EPI_ISL_546524\|2020-0 | ···················· | ···················· | ···················· | ···················· | ···················· | ···················· | |
| I | 1149 hCoV-19/USA/TX-HMH-7124/2020\|EPI_ISL_546525\|2020-0 | ···················· | ···················· | ···················· | ···················· | ···················· | ···················· | |
| I | 1150 hCoV-19/USA/TX-HMH-7125/2020\|EPI_ISL_546526\|2020-0 | ···················· | ···················· | ···················· | ···················· | ···················· | ···················· | |
| I | 1151 hCoV-19/USA/TX-HMH-7126/2020\|EPI_ISL_546527\|2020-0 | ···················· | ···················· | ···················· | ···················· | ···················· | ···················· | |
| I | 1152 hCoV-19/USA/TX-HMH-7129/2020\|EPI_ISL_546529\|2020-0 | ···················· | ···················· | ···················· | ···················· | ···················· | ···················· | |
| I | 1153 hCoV-19/USA/TX-HMH-7131/2020\|EPI_ISL_546531\|2020-0 | ···················· | ·············A······ | ···················· | ···················· | ···················· | ···················· | |
| I | 1157 hCoV-19/USA/TX-HMH-7139/2020\|EPI_ISL_546538\|2020-0 | ···················· | ···················· | ···················· | ···················· | ···················· | ···················· | |
| I | 1159 hCoV-19/USA/TX-HMH-7143/2020\|EPI_ISL_546541\|2020-0 | ···················· | ···················· | ···················· | ···················· | ···················· | ···················· | |
| I | 1163 hCoV-19/USA/TX-HMH-7149/2020\|EPI_ISL_546547\|2020-0 | ···················· | ···················· | ···················· | ···················· | ···················· | ···················· | |
| I | 1164 hCoV-19/USA/TX-HMH-5768/2020\|EPI_ISL_547000\|2020-0 | ···················· | ·············A······ | ···················· | ···················· | ··············NN···· | ···················· | |
| I | 1165 hCoV-19/USA/TX-HMH-577/2020\|EPI_ISL_547001\|2020-04 | ···················· | ···················· | ···················· | ···················· | ···················· | ···················· | |
| I | 1166 hCoV-19/USA/TX-HMH-5770/2020\|EPI_ISL_547002\|2020-0 | ···················· | ···················· | ···················· | ···················· | ···················· | ···················· | |
| I | 1167 hCoV-19/USA/TX-HMH-5771/2020\|EPI_ISL_547003\|2020-0 | ···················· | ·············A······ | ···················· | ···················· | ···················· | ···················· | |
| I | 1168 hCoV-19/USA/TX-HMH-5772/2020\|EPI_ISL_547004\|2020-0 | ···················· | ···················· | ···················· | ···················· | ···················· | ···················· | |
| I | 1169 hCoV-19/USA/TX-HMH-5776/2020\|EPI_ISL_547007\|2020-0 | ···················· | ·A·················· | ···················· | ···················· | ···················· | ···················· | |
| I | 1170 hCoV-19/USA/TX-HMH-5778/2020\|EPI_ISL_547008\|2020-0 | ···················· | ···················· | ···················· | ···················· | ···················· | ···················· | |
| I | 1171 hCoV-19/USA/TX-HMH-5781/2020\|EPI_ISL_547009\|2020-0 | ···················· | ···················· | ···················· | ···················· | ···················· | ···················· | |
| I | 1172 hCoV-19/USA/TX-HMH-5782/2020\|EPI_ISL_547010\|2020-0 | ···················· | ···················· | ···················· | ···················· | ···················· | ···················· | |
| I | 1173 hCoV-19/USA/TX-HMH-5783/2020\|EPI_ISL_547011\|2020-0 | ···················· | ···················· | ···················· | ···················· | ···················· | ···················· | |
| I | 1175 hCoV-19/USA/TX-HMH-5786/2020\|EPI_ISL_547013\|2020-0 | ···················· | ···················· | ···················· | ···················· | ···················· | ···················· | |
| I | 1176 hCoV-19/USA/TX-HMH-5787/2020\|EPI_ISL_547014\|2020-0 | ···················· | ···················· | ···················· | ···················· | ···················· | ···················· | |
| I | 1177 hCoV-19/USA/TX-HMH-5788/2020\|EPI_ISL_547015\|2020-0 | ···················· | ···················· | ···················· | ···················· | ···················· | ···················· | |
| I | 1178 hCoV-19/USA/TX-HMH-5789/2020\|EPI_ISL_547016\|2020-0 | ···················· | ···················· | ···················· | ···················· | ···················· | ···················· | |
| I | 1183 hCoV-19/USA/TX-HMH-5880/2020\|EPI_ISL_547083\|2020-0 | ···················· | ·A·················· | ···················· | ···················· | ···················· | ······NN············ | |
| I | 1186 hCoV-19/USA/TX-HMH-5886/2020\|EPI_ISL_547087\|2020-0 | ···················· | ···················· | ···················· | ···················· | ···················· | ···················· | |
| I | 1187 hCoV-19/USA/TX-HMH-5890/2020\|EPI_ISL_547088\|2020-0 | ···················· | ·A·················· | ···················· | ···················· | ···················· | ···················· | |
| I | 1189 hCoV-19/USA/TX-HMH-5896/2020\|EPI_ISL_547092\|2020-0 | ···················· | ···················· | ···················· | ···················· | ···················· | ···················· | |
| I | 1191 hCoV-19/USA/TX-HMH-5900/2020\|EPI_ISL_547095\|2020-0 | ···················· | ···················· | ···················· | ···················· | ···················· | ···················· | |
| I | 1192 hCoV-19/USA/TX-HMH-5901/2020\|EPI_ISL_547096\|2020-0 | ···················· | ···················· | ···················· | ···················· | ···················· | ···················· | |
| I | 1193 hCoV-19/USA/TX-HMH-5902/2020\|EPI_ISL_547097\|2020-0 | ···················· | ···················· | ···················· | ···················· | ···················· | ···················· | |
| I | 1195 hCoV-19/USA/WI-GMF-19268/2020\|EPI_ISL_547802\|2020- | ···················· | ···················· | ···················· | ···················· | ···················· | ···················· | |
| I | 1196 hCoV-19/USA/WI-GMF-19271/2020\|EPI_ISL_547803\|2020- | ···················· | ···················· | ···················· | ···················· | ···················· | ···················· | |
| I | 1197 hCoV-19/USA/WI-GMF-19273/2020\|EPI_ISL_547804\|2020- | ···················· | ···················· | ···················· | ···················· | ···················· | ···················· | |
| I | 1198 hCoV-19/USA/WI-GMF-19275/2020\|EPI_ISL_547805\|2020- | ···················· | ···················· | ···················· | ···················· | ···················· | ···················· | |
| I | 1199 hCoV-19/USA/WI-GMF-19279/2020\|EPI_ISL_547806\|2020- | ···················· | ···················· | ···················· | ···················· | ···················· | ···················· | |
| I | 1200 hCoV-19/USA/WI-GMF-19309/2020\|EPI_ISL_547807\|2020- | ···················· | ···················· | ···················· | ···················· | ···················· | ···················· | |
| I | 1201 hCoV-19/USA/WI-GMF-19443/2020\|EPI_ISL_547808\|2020- | ···················· | ···················· | ···················· | ···················· | ···················· | ···················· | |
| I | 1202 hCoV-19/USA/WI-GMF-19457/2020\|EPI_ISL_547809\|2020- | ···················· | ···················· | ···················· | ···················· | ···················· | ···················· | |
| I | 1203 hCoV-19/USA/WI-GMF-19466/2020\|EPI_ISL_547810\|2020- | ···················· | ···················· | ···················· | ···················· | ···················· | ···············A···· | |
| I | 1204 hCoV-19/USA/WI-GMF-19468/2020\|EPI_ISL_547811\|2020- | ···················· | ···················· | ···················· | ···················· | ···················· | ···················· | |
| I | 1205 hCoV-19/USA/WI-GMF-19488/2020\|EPI_ISL_547812\|2020- | ···················· | ···················· | ···················· | ···················· | ···················· | ···················· | |
| I | 1206 hCoV-19/USA/WI-GMF-19490/2020\|EPI_ISL_547813\|2020- | ···················· | ···················· | ···················· | ···················· | ···················· | ···················· | |
| I | 1207 hCoV-19/USA/WI-GMF-19492/2020\|EPI_ISL_547814\|2020- | ···················· | ···················· | ···················· | ···················· | ···················· | ···················· | |
| I | 1208 hCoV-19/USA/WI-GMF-19494/2020\|EPI_ISL_547815\|2020- | ···················· | ···················· | ···················· | ···················· | ···················· | ···················· | |
| I | 1209 hCoV-19/USA/WI-GMF-19500/2020\|EPI_ISL_547816\|2020- | ···················· | ···················· | ···················· | ···················· | ···················· | ···················· | |
| I | 1210 hCoV-19/USA/WI-GMF-19502/2020\|EPI_ISL_547817\|2020- | ···················· | ·············A······ | ···················· | ···················· | ···················· | ···················· | |
| I | 1211 hCoV-19/USA/WI-GMF-19523/2020\|EPI_ISL_547818\|2020- | ···················· | ···················· | ···················· | ···················· | ···················· | ···················· | |
| I | 1212 hCoV-19/USA/WI-GMF-19557/2020\|EPI_ISL_547819\|2020- | ···················· | ···················· | ···················· | ···················· | ···················· | ···················· | |
| I | 1213 hCoV-19/USA/WI-GMF-19558/2020\|EPI_ISL_547820\|2020- | ···················· | ···················· | ···················· | ···················· | ···················· | ···················· | |
| I | 1214 hCoV-19/USA/WI-GMF-19560/2020\|EPI_ISL_547821\|2020- | ···················· | ···················· | ···················· | ···················· | ···················· | ···················· | |
| I | 1215 hCoV-19/USA/WI-GMF-19565/2020\|EPI_ISL_547822\|2020- | ···················· | ···················· | ···················· | ···················· | ···················· | ···················· | |
| I | 1216 hCoV-19/USA/WI-GMF-19580/2020\|EPI_ISL_547823\|2020- | ···················· | ···················· | ···················· | ···················· | ···················· | ···················· | |
| I | 1217 hCoV-19/USA/WI-GMF-19586/2020\|EPI_ISL_547824\|2020- | ···················· | ···················· | ···················· | ···················· | ···················· | ···················· | |
| I | 1218 hCoV-19/USA/WI-GMF-19593/2020\|EPI_ISL_547825\|2020- | ···················· | ···················· | ···················· | ···················· | ···················· | ···················· | |
| I | 1219 hCoV-19/USA/WI-GMF-19596/2020\|EPI_ISL_547826\|2020- | ···················· | ···················· | ···················· | ···················· | ···················· | ···················· | |
| I | 1220 hCoV-19/USA/WI-GMF-19598/2020\|EPI_ISL_547827\|2020- | ···················· | ···················· | ···················· | ···················· | ···················· | ···················· | |
| I | 1221 hCoV-19/USA/WI-GMF-19599/2020\|EPI_ISL_547828\|2020- | ···················· | ···················· | ···················· | ···················· | ···················· | ···················· | |

| | | | | | | | | |
|---|---|---|---|---|---|---|---|---|
| I | 1222 hCoV-19/USA/WI-GMF-19600/2020\|EPI_ISL_547829\|2020- | ···················· | ···················· | ····················· | ···················· | ···················· | ···················· | |
| I | 1223 hCoV-19/USA/WI-GMF-19604/2020\|EPI_ISL_547831\|2020- | ···················· | ···················· | ····················· | ···················· | ···················· | ···················· | |
| I | 1224 hCoV-19/USA/WI-GMF-19628/2020\|EPI_ISL_547833\|2020- | ···················· | ···················· | ····················· | ···················· | ···················· | ···················· | |
| I | 1225 hCoV-19/USA/WI-GMF-19656/2020\|EPI_ISL_547834\|2020- | ···················· | ···················· | ····················· | ···················· | ···················· | ···················· | |
| I | 1226 hCoV-19/USA/WI-GMF-19668/2020\|EPI_ISL_547835\|2020- | ···················· | ···················· | ····················· | ···················· | ···················· | ···················· | |
| I | 1227 hCoV-19/USA/WI-GMF-19707/2020\|EPI_ISL_547836\|2020- | ···················· | ···················· | ····················· | ···················· | ···················· | ···············A···· | |
| I | 1228 hCoV-19/USA/WI-GMF-19708/2020\|EPI_ISL_547837\|2020- | ···················· | ···················· | ····················· | ···················· | ···················· | ···················· | |
| I | 1229 hCoV-19/USA/WI-GMF-19709/2020\|EPI_ISL_547838\|2020- | ···················· | ···················· | ····················· | ···················· | ···················· | ···················· | |
| I | 1230 hCoV-19/USA/WI-GMF-19714/2020\|EPI_ISL_547839\|2020- | ···················· | ···················· | ····················· | ···················· | ···················· | ···················· | |
| I | 1231 hCoV-19/USA/WI-GMF-19716/2020\|EPI_ISL_547840\|2020- | ···················· | ···················· | ····················· | ···················· | ···················· | ···················· | |
| I | 1232 hCoV-19/USA/WI-GMF-19729/2020\|EPI_ISL_547841\|2020- | ···················· | ···················· | ····················· | ···················· | ···················· | ···················· | |
| I | 1233 hCoV-19/USA/WI-GMF-19732/2020\|EPI_ISL_547842\|2020- | ···················· | ···················· | ····················· | ···················· | ···················· | ···············A···· | |
| I | 1234 hCoV-19/USA/WI-GMF-19743/2020\|EPI_ISL_547843\|2020- | ···················· | ···················· | ····················· | ···················· | ···················· | ···················· | |
| I | 1235 hCoV-19/USA/WI-GMF-19745/2020\|EPI_ISL_547844\|2020- | ···················· | ···················· | ····················· | ···················· | ···················· | ···················· | |
| I | 1236 hCoV-19/USA/WI-GMF-19746/2020\|EPI_ISL_547845\|2020- | ···················· | ···················· | ····················· | ···················· | ···················· | ···············A···· | |
| I | 1237 hCoV-19/USA/WI-GMF-19787/2020\|EPI_ISL_547846\|2020- | ···················· | ···················· | ····················· | ···················· | ···················· | ···················· | |
| I | 1238 hCoV-19/USA/WI-GMF-19935/2020\|EPI_ISL_547847\|2020- | ···················· | ···················· | ····················· | ···················· | ···················· | ···················· | |
| I | 1239 hCoV-19/USA/WI-GMF-19968/2020\|EPI_ISL_547848\|2020- | ···················· | ···················· | ····················· | ···················· | ···················· | ···················· | |
| I | 1240 hCoV-19/USA/WI-GMF-20111/2020\|EPI_ISL_547849\|2020- | ···················· | ···················· | ····················· | ···················· | ···················· | ···················· | |
| I | 1241 hCoV-19/USA/WI-GMF-20119/2020\|EPI_ISL_547850\|2020- | ···················· | ···················· | ····················· | ···················· | ···················· | ···················· | |
| I | 1242 hCoV-19/USA/WI-GMF-20122/2020\|EPI_ISL_547851\|2020- | ···················· | ···················· | ····················· | ···················· | ···················· | ···················· | |
| I | 1243 hCoV-19/USA/WI-GMF-20149/2020\|EPI_ISL_547853\|2020- | ···················· | ···················· | ····················· | ···················· | ···················· | ···················· | |
| I | 1244 hCoV-19/USA/WI-GMF-20172/2020\|EPI_ISL_547855\|2020- | ···················· | ···················· | ····················· | ···················· | ···················· | ···················· | |
| I | 1245 hCoV-19/USA/WI-GMF-20177/2020\|EPI_ISL_547856\|2020- | ···················· | ···················· | ····················· | ···················· | ···················· | ···················· | |
| I | 1246 hCoV-19/USA/IA-GMF-20217/2020\|EPI_ISL_547857\|2020- | ···················· | ·A·················· | ····················· | ···················· | ···················· | ···················· | |
| I | 1247 hCoV-19/USA/WI-GMF-20228/2020\|EPI_ISL_547858\|2020- | ···················· | ···················· | ····················· | ···················· | ···················· | ···················· | |
| I | 1248 hCoV-19/USA/WI-GMF-20271/2020\|EPI_ISL_547859\|2020- | ···················· | ···················· | ····················· | ···················· | ···················· | ···················· | |
| I | 1249 hCoV-19/USA/WI-GMF-20273/2020\|EPI_ISL_547860\|2020- | ···················· | ···················· | ····················· | ···················· | ···················· | ···················· | |
| I | 1250 hCoV-19/USA/WI-GMF-20276/2020\|EPI_ISL_547861\|2020- | ···················· | ···················· | ····················· | ···················· | ···················· | ···················· | |
| I | 1251 hCoV-19/USA/WI-GMF-20295/2020\|EPI_ISL_547862\|2020- | ···················· | ···················· | ····················· | ···················· | ···················· | ···················· | |
| I | 1252 hCoV-19/USA/WI-GMF-20333/2020\|EPI_ISL_547863\|2020- | ···················· | ···················· | ····················· | ···················· | ···················· | ···················· | |
| I | 1253 hCoV-19/USA/WI-GMF-20334/2020\|EPI_ISL_547864\|2020- | ···················· | ···················· | ····················· | ···················· | ···················· | ···················· | |
| I | 1254 hCoV-19/USA/WI-GMF-20378/2020\|EPI_ISL_547865\|2020- | ···················· | ···················· | ····················· | ···················· | ···················· | ···················· | |
| I | 1255 hCoV-19/USA/WI-GMF-20389/2020\|EPI_ISL_547866\|2020- | ···················· | ···················· | ····················· | ···················· | ···················· | ···················· | |
| I | 1256 hCoV-19/USA/WI-GMF-20407/2020\|EPI_ISL_547867\|2020- | ···················· | ···················· | ····················· | ···················· | ···················· | ···············A···· | |
| I | 1257 hCoV-19/USA/WI-GMF-20442/2020\|EPI_ISL_547868\|2020- | ···················· | ···················· | ····················· | ···················· | ···················· | ···················· | |
| I | 1258 hCoV-19/USA/WI-GMF-20447/2020\|EPI_ISL_547869\|2020- | ···················· | ···················· | ····················· | ···················· | ···················· | ···················· | |
| I | 1259 hCoV-19/USA/CA-CZB-3275/2020\|EPI_ISL_548319\|2020-0 | ···················· | ···················· | ····················· | ···················· | ···················· | ···················· | |
| I | 1260 hCoV-19/USA/CA-CZB-3309/2020\|EPI_ISL_548322\|2020-0 | ···················· | ·A·················· | ····················· | ···················· | ···················· | ···················· | |
| I | 1261 hCoV-19/USA/CA-CZB-3365/2020\|EPI_ISL_548323\|2020-0 | ···················· | ·A·················· | ····················· | ···················· | ···················· | ···················· | |
| I | 1262 hCoV-19/USA/CA-CZB-3321/2020\|EPI_ISL_548325\|2020-0 | ···················· | ·A·················· | ····················· | ···················· | ···················· | ···················· | |
| I | 1263 hCoV-19/USA/CA-CZB-3368/2020\|EPI_ISL_548326\|2020-0 | ···················· | ···················· | ····················· | ···················· | ···················· | ···················· | |
| I | 1264 hCoV-19/USA/CA-CZB-3294/2020\|EPI_ISL_548327\|2020-0 | ···················· | ···················· | ····················· | ···················· | ···················· | ···················· | |
| I | 1265 hCoV-19/USA/CA-CZB-3342/2020\|EPI_ISL_548328\|2020-0 | ···················· | ···················· | ····················· | ···················· | ···················· | ···················· | |
| I | 1266 hCoV-19/USA/CA-CZB-3308/2020\|EPI_ISL_548329\|2020-0 | ···················· | ···················· | ····················· | ···················· | ···················· | ···················· | |
| I | 1267 hCoV-19/USA/CA-CZB-3363/2020\|EPI_ISL_548332\|2020-0 | ···················· | ···················· | ····················· | ···················· | ···················· | ···················· | |
| I | 1268 hCoV-19/USA/CA-CZB-3340/2020\|EPI_ISL_548334\|2020-0 | ···················· | ·A·················· | ····················· | ···················· | ···················· | ···················· | |
| I | 1269 hCoV-19/USA/CA-CZB-3304/2020\|EPI_ISL_548336\|2020-0 | ···················· | ···················· | ····················· | ···················· | ···················· | ···················· | |
| I | 1270 hCoV-19/USA/CA-CZB-3620/2020\|EPI_ISL_548337\|2020-0 | ···················· | ·A·················· | ····················· | ···················· | ···················· | ···················· | |
| I | 1271 hCoV-19/USA/CA-CZB-3621/2020\|EPI_ISL_548338\|2020-0 | ···················· | ·A·················· | ····················· | ···················· | ···················· | ···················· | |
| I | 1272 hCoV-19/USA/CA-CZB-3616/2020\|EPI_ISL_548339\|2020-0 | ···················· | ·A·················· | ····················· | ···················· | ···················· | ···················· | |
| I | 1273 hCoV-19/USA/CA-CZB-3622/2020\|EPI_ISL_548340\|2020-0 | ···················· | ·A·················· | ····················· | ···················· | ···················· | ···················· | |
| I | 1274 hCoV-19/USA/CA-CZB-3619/2020\|EPI_ISL_548341\|2020-0 | ···················· | ·A·················· | ····················· | ···················· | ···················· | ···················· | |
| I | 1275 hCoV-19/USA/CA-CZB-3618/2020\|EPI_ISL_548342\|2020-0 | ···················· | ·A·················· | ····················· | ···················· | ···················· | ···················· | |
| I | 1276 hCoV-19/USA/CA-CZB-3615/2020\|EPI_ISL_548343\|2020-0 | ···················· | ·A·················· | ····················· | ···················· | ···················· | ···················· | |
| I | 1277 hCoV-19/USA/CA-CZB-3617/2020\|EPI_ISL_548344\|2020-0 | ···················· | ·A·················· | ····················· | ···················· | ···················· | ···················· | |
| I | 1278 hCoV-19/USA/CA-CZB-3623/2020\|EPI_ISL_548345\|2020-0 | ···················· | ···················· | ····················· | ···················· | ···················· | ···················· | |
| I | 1279 hCoV-19/USA/CA-CZB-3640/2020\|EPI_ISL_548346\|2020-0 | ···················· | ···················· | ····················· | ···················· | ···················· | ···················· | |
| I | 1280 hCoV-19/USA/CA-CZB-3634/2020\|EPI_ISL_548347\|2020-0 | ···················· | ···················· | ····················· | ···················· | ···················· | ···················· | |
| I | 1281 hCoV-19/USA/TX-HMH-5275/2020\|EPI_ISL_544645\|2020-0 | ···················· | ···················· | ····················· | ···················· | ···················· | ···················· | |
| I | 1282 hCoV-19/USA/TX-HMH-5277/2020\|EPI_ISL_544646\|2020-0 | ···················· | ···················· | ····················· | ···················· | ··············N····· | NN·················· | |
| I | 1283 hCoV-19/USA/TX-HMH-5282/2020\|EPI_ISL_544651\|2020-0 | ···················· | ···················· | ····················· | ···················· | ···················· | ···················· | |
| I | 1287 hCoV-19/USA/TX-HMH-2068/2020\|EPI_ISL_545101\|2020-0 | ···················· | ···················· | ····················· | ···················· | ···················· | ···················· | |
| I | 1288 hCoV-19/USA/TX-HMH-2069/2020\|EPI_ISL_545102\|2020-0 | ···················· | ···················· | ····················· | ···················· | ···················· | ···················· | |
| I | 1289 hCoV-19/USA/TX-HMH-2073/2020\|EPI_ISL_545103\|2020-0 | ···················· | ···················· | ····················· | ···················· | ···················· | ···················· | |
| I | 1290 hCoV-19/USA/TX-HMH-2077/2020\|EPI_ISL_545106\|2020-0 | ···················· | ···················· | ····················· | ···················· | ···················· | ···················· | |
| I | 1291 hCoV-19/USA/TX-HMH-2079/2020\|EPI_ISL_545108\|2020-0 | ···················· | ···················· | ····················· | ···················· | ···················· | ···················· | |
| I | 1292 hCoV-19/USA/TX-HMH-2080/2020\|EPI_ISL_545109\|2020-0 | ···················· | ···················· | ····················· | ···················· | ···················· | ···················· | |
| I | 1293 hCoV-19/USA/TX-HMH-2081/2020\|EPI_ISL_545110\|2020-0 | ···················· | ···················· | ····················· | ···················· | ···················· | ···················· | |
| I | 1294 hCoV-19/USA/TX-HMH-2082/2020\|EPI_ISL_545111\|2020-0 | ···················· | ···················· | ····················· | ···················· | ···················· | ···················· | |
| I | 1295 hCoV-19/USA/TX-HMH-2087/2020\|EPI_ISL_545112\|2020-0 | ···················· | ···················· | ····················· | ···················· | ···················· | ···················· | |
| I | 1296 hCoV-19/USA/TX-HMH-2088/2020\|EPI_ISL_545113\|2020-0 | ···················· | ···················· | ····················· | ···················· | ···················· | ···················· | |
| I | 1297 hCoV-19/USA/TX-HMH-2090/2020\|EPI_ISL_545114\|2020-0 | ···················· | ···················· | ····················· | ···················· | ···················· | ···················· | |
| I | 1298 hCoV-19/USA/TX-HMH-2092/2020\|EPI_ISL_545115\|2020-0 | ···················· | ···················· | ····················· | ···················· | ···················· | ···················· | |
| I | 1299 hCoV-19/USA/TX-HMH-2093/2020\|EPI_ISL_545116\|2020-0 | ···················· | ···················· | ····················· | ···················· | ···················· | ···················· | |
| I | 1300 hCoV-19/USA/TX-HMH-2094/2020\|EPI_ISL_545117\|2020-0 | ···················· | ···················· | ····················· | ···················· | ···················· | ···················· | |
| I | 1301 hCoV-19/USA/TX-HMH-2095/2020\|EPI_ISL_545118\|2020-0 | ···················· | ···················· | ····················· | ···················· | ···················· | NN·····N············ | 1 |
| I | 1302 hCoV-19/USA/TX-HMH-2098/2020\|EPI_ISL_545119\|2020-0 | ···················· | ···················· | ····················· | ···················· | ···················· | ···················· | |
| I | 1303 hCoV-19/USA/TX-HMH-2100/2020\|EPI_ISL_545120\|2020-0 | ···················· | ··············C····· | ····················· | ···················· | ···················· | ···················· | |
| I | 1304 hCoV-19/USA/TX-HMH-2101/2020\|EPI_ISL_545121\|2020-0 | ···················· | ···················· | ····················· | ···················· | ···················· | ···················· | |

| | | | | | | | | | |
|---|---|---|---|---|---|---|---|---|---|
| I | 1305 | hCoV-19/USA/TX-HMH-2102/2020\|EPI_ISL_545122\|2020-0 | ···················· | ···················· | ···················· | ···················· | ···················· | ···················· | |
| I | 1306 | hCoV-19/USA/TX-HMH-2103/2020\|EPI_ISL_545123\|2020-0 | ···················· | ······A············· | ···················· | ···················· | ···················· | ···················· | |
| I | 1307 | hCoV-19/USA/TX-HMH-2107/2020\|EPI_ISL_545125\|2020-0 | ···················· | ···················· | ···················· | ···················· | ···················· | ···················· | |
| I | 1308 | hCoV-19/USA/TX-HMH-2109/2020\|EPI_ISL_545126\|2020-0 | ···················· | ···················· | ···················· | ···················· | ···················· | ···················· | |
| I | 1309 | hCoV-19/USA/TX-HMH-2113/2020\|EPI_ISL_545127\|2020-0 | ···················· | ···················· | ···················· | ···················· | ···················· | ···················· | |
| I | 1310 | hCoV-19/USA/TX-HMH-2115/2020\|EPI_ISL_545128\|2020-0 | ···················· | ···················· | ···················· | ···················· | ···················· | ···················· | |
| I | 1311 | hCoV-19/USA/TX-HMH-2118/2020\|EPI_ISL_545129\|2020-0 | ···················· | ··············C····· | ···················· | ···················· | ···················· | ···················· | |
| I | 1312 | hCoV-19/USA/TX-HMH-2119/2020\|EPI_ISL_545130\|2020-0 | ···················· | ···················· | ···················· | ···················· | ···················· | ···················· | |
| I | 1313 | hCoV-19/USA/TX-HMH-2121/2020\|EPI_ISL_545131\|2020-0 | ···················· | ···················· | ···················· | ···················· | ···················· | ···················· | |
| I | 1314 | hCoV-19/USA/TX-HMH-2123/2020\|EPI_ISL_545132\|2020-0 | ···················· | ···················· | ···················· | ···················· | ···················· | ···················· | |
| I | 1315 | hCoV-19/USA/TX-HMH-2124/2020\|EPI_ISL_545133\|2020-0 | ···················· | ···················· | ···················· | ···················· | ···················· | ···················· | |
| I | 1316 | hCoV-19/USA/TX-HMH-2125/2020\|EPI_ISL_545134\|2020-0 | ···················· | ···················· | ···················· | ···················· | ···················· | ···················· | |
| I | 1317 | hCoV-19/USA/TX-HMH-2129/2020\|EPI_ISL_545135\|2020-0 | ···················· | ···················· | ···················· | ···················· | ···················· | ···················· | |
| I | 1318 | hCoV-19/USA/TX-HMH-2131/2020\|EPI_ISL_545136\|2020-0 | ···················· | ···················· | ···················· | ···················· | ···················· | ···················· | |
| I | 1319 | hCoV-19/USA/TX-HMH-2133/2020\|EPI_ISL_545137\|2020-0 | ···················· | ···················· | ···················· | ···················· | ···················· | ···················· | |
| I | 1320 | hCoV-19/USA/TX-HMH-2135/2020\|EPI_ISL_545138\|2020-0 | ···················· | ···················· | ···················· | ···················· | ···················· | ···················· | |
| I | 1321 | hCoV-19/USA/TX-HMH-2136/2020\|EPI_ISL_545139\|2020-0 | ···················· | ···················· | ···················· | ···················· | ···················· | ···················· | |
| I | 1322 | hCoV-19/USA/TX-HMH-2137/2020\|EPI_ISL_545140\|2020-0 | ···················· | ···················· | ···················· | ···················· | ···················· | ···················· | |
| I | 1323 | hCoV-19/USA/TX-HMH-2142/2020\|EPI_ISL_545142\|2020-0 | ···················· | ······A············· | ···················· | ···················· | ···················· | ···················· | |
| I | 1324 | hCoV-19/USA/TX-HMH-2145/2020\|EPI_ISL_545143\|2020-0 | ···················· | ···················· | ···················· | ···················· | ···················· | ···················· | |
| I | 1325 | hCoV-19/USA/TX-HMH-2148/2020\|EPI_ISL_545144\|2020-0 | ···················· | ······A············· | ···················· | ···················· | ···················· | ···················· | |
| I | 1326 | hCoV-19/USA/TX-HMH-2149/2020\|EPI_ISL_545145\|2020-0 | ···················· | ···················· | ···················· | ···················· | ···················· | ···················· | |
| I | 1327 | hCoV-19/USA/TX-HMH-2150/2020\|EPI_ISL_545146\|2020-0 | ···················· | ···················· | ···················· | ···················· | ···················· | ···················· | |
| I | 1328 | hCoV-19/USA/TX-HMH-2153/2020\|EPI_ISL_545147\|2020-0 | ···················· | ···················· | ···················· | ···················· | ···················· | ···················· | |
| I | 1329 | hCoV-19/USA/TX-HMH-2157/2020\|EPI_ISL_545149\|2020-0 | ···················· | ··············C····· | ···················· | ···················· | ···················· | ···················· | |
| I | 1330 | hCoV-19/USA/TX-HMH-2158/2020\|EPI_ISL_545150\|2020-0 | ···················· | ···················· | ···················· | ···················· | ···················· | ···················· | |
| I | 1331 | hCoV-19/USA/TX-HMH-2161/2020\|EPI_ISL_545152\|2020-0 | ···················· | ···················· | ···················· | ···················· | ···················· | ···················· | |
| I | 1332 | hCoV-19/USA/TX-HMH-2162/2020\|EPI_ISL_545153\|2020-0 | ···················· | ···················· | ···················· | ···················· | ···················· | ···················· | |
| I | 1333 | hCoV-19/USA/TX-HMH-2165/2020\|EPI_ISL_545155\|2020-0 | ···················· | ···················· | ···················· | ···················· | ··············N·N··· | N··N················ | |
| I | 1334 | hCoV-19/USA/TX-HMH-2166/2020\|EPI_ISL_545156\|2020-0 | ···················· | ···················· | ···················· | ···················· | ···················· | ···················· | |
| I | 1335 | hCoV-19/USA/TX-HMH-2168/2020\|EPI_ISL_545157\|2020-0 | ···················· | ···················· | ···················· | ···················· | ···················· | ···················· | |
| I | 1336 | hCoV-19/USA/TX-HMH-2169/2020\|EPI_ISL_545158\|2020-0 | ···················· | ···················· | ···················· | ···················· | ···················· | ···················· | |
| I | 1337 | hCoV-19/USA/TX-HMH-2170/2020\|EPI_ISL_545159\|2020-0 | ···················· | ···················· | ···················· | ···················· | ···················· | ···················· | |
| I | 1338 | hCoV-19/USA/TX-HMH-2171/2020\|EPI_ISL_545160\|2020-0 | ···················· | ···················· | ···················· | ···················· | ···················· | ···················· | |
| I | 1339 | hCoV-19/USA/TX-HMH-2173/2020\|EPI_ISL_545161\|2020-0 | ···················· | ···················· | ···················· | ···················· | ···················· | ···················· | |
| I | 1340 | hCoV-19/USA/TX-HMH-2174/2020\|EPI_ISL_545162\|2020-0 | ···················· | ······A············· | ···················· | ···················· | ···················· | ···················· | |
| I | 1341 | hCoV-19/USA/TX-HMH-2175/2020\|EPI_ISL_545163\|2020-0 | ···················· | ···················· | ···················· | ···················· | ···················· | ···················· | |
| I | 1342 | hCoV-19/USA/TX-HMH-2177/2020\|EPI_ISL_545164\|2020-0 | ···················· | ···················· | ···················· | ···················· | ···················· | ···················· | |
| I | 1343 | hCoV-19/USA/TX-HMH-2178/2020\|EPI_ISL_545165\|2020-0 | ···················· | ···················· | ···················· | ···················· | ···················· | ···················· | |
| I | 1344 | hCoV-19/USA/TX-HMH-2179/2020\|EPI_ISL_545166\|2020-0 | ···················· | ··············C····· | ···················· | ···················· | ···················· | NN·················· | |
| I | 1346 | hCoV-19/USA/TX-HMH-2185/2020\|EPI_ISL_545169\|2020-0 | ···················· | ···················· | ···················· | ···················· | ···················· | ···················· | |
| I | 1347 | hCoV-19/USA/TX-HMH-2186/2020\|EPI_ISL_545170\|2020-0 | ···················· | ······A············· | ···················· | ···················· | ···················· | ···················· | |
| I | 1348 | hCoV-19/USA/TX-HMH-2189/2020\|EPI_ISL_545171\|2020-0 | ···················· | ···················· | ···················· | ···················· | ···················· | ···················· | |
| I | 1349 | hCoV-19/USA/TX-HMH-2193/2020\|EPI_ISL_545172\|2020-0 | ···················· | ···················· | ···················· | ···················· | ···················· | ···················· | |
| I | 1350 | hCoV-19/USA/TX-HMH-2195/2020\|EPI_ISL_545173\|2020-0 | ···················· | ······A············· | ···················· | ···················· | ···················· | ···················· | |
| I | 1352 | hCoV-19/USA/TX-HMH-2198/2020\|EPI_ISL_545176\|2020-0 | ···················· | ···················· | ···················· | ···················· | ···················· | ···················· | |
| I | 1353 | hCoV-19/USA/TX-HMH-2200/2020\|EPI_ISL_545177\|2020-0 | ···················· | ···················· | ···················· | ···················· | ···················· | ···················· | |
| I | 1354 | hCoV-19/USA/TX-HMH-2201/2020\|EPI_ISL_545178\|2020-0 | ···················· | ···················· | ···················· | ···················· | ···················· | ···················· | |
| I | 1355 | hCoV-19/USA/TX-HMH-2202/2020\|EPI_ISL_545179\|2020-0 | ···················· | ···················· | ···················· | ···················· | ···················· | ···················· | |
| I | 1357 | hCoV-19/USA/TX-HMH-6406/2020\|EPI_ISL_545977\|2020-0 | ···················· | ···················· | ···················· | ···················· | ···················· | ···················· | |
| I | 1358 | hCoV-19/USA/TX-HMH-6411/2020\|EPI_ISL_545981\|2020-0 | ···················· | ···················· | ···················· | ···················· | ···················· | ···················· | |
| I | 1359 | hCoV-19/USA/TX-HMH-6412/2020\|EPI_ISL_545982\|2020-0 | ···················· | ···················· | ···················· | ···················· | ···················· | ···················· | |
| I | 1360 | hCoV-19/USA/TX-HMH-6414/2020\|EPI_ISL_545983\|2020-0 | ···················· | ···················· | ···················· | ···················· | ···················· | ···················· | |
| I | 1361 | hCoV-19/USA/TX-HMH-6419/2020\|EPI_ISL_545985\|2020-0 | ···················· | ···················· | ···················· | ···················· | ··············N····· | N··················· | |
| I | 1368 | hCoV-19/Czech Republic/NRL-7950/2020\|EPI_ISL_54643 | ···················· | ···················· | ···················· | ···················· | ···················· | ···················· | |
| I | 1369 | hCoV-19/USA/TX-HMH-7018/2020\|EPI_ISL_546437\|2020-0 | ···················· | ···················· | ···················· | ···················· | ···················· | ···················· | |
| I | 1370 | hCoV-19/USA/TX-HMH-7019/2020\|EPI_ISL_546438\|2020-0 | ···················· | ·············A······ | ···················· | ···················· | ···················· | ···················· | |
| I | 1372 | hCoV-19/USA/TX-HMH-7020/2020\|EPI_ISL_546440\|2020-0 | ···················· | ···················· | ···················· | ···················· | ······T············· | ···················· | |
| I | 1373 | hCoV-19/USA/TX-HMH-7024/2020\|EPI_ISL_546443\|2020-0 | ···················· | ···················· | ···················· | ···················· | ···················· | ···················· | |
| I | 1374 | hCoV-19/USA/TX-HMH-7025/2020\|EPI_ISL_546444\|2020-0 | ···················· | ·A·················· | ···················· | ···················· | ··············N····· | ···················· | |
| I | 1376 | hCoV-19/USA/TX-HMH-7029/2020\|EPI_ISL_546447\|2020-0 | ···················· | ···················· | ···················· | ···················· | ···················· | ···················· | |
| I | 1377 | hCoV-19/USA/TX-HMH-7030/2020\|EPI_ISL_546448\|2020-0 | ···················· | ···················· | ···················· | ···················· | ···················· | ···················· | |
| I | 1379 | hCoV-19/USA/TX-HMH-7032/2020\|EPI_ISL_546450\|2020-0 | ···················· | ···················· | ···················· | ···················· | ···················· | ···················· | |
| I | 1380 | hCoV-19/USA/TX-HMH-7033/2020\|EPI_ISL_546451\|2020-0 | ···················· | ···················· | ···················· | ···················· | ···················· | ···················· | |
| I | 1381 | hCoV-19/USA/TX-HMH-7036/2020\|EPI_ISL_546454\|2020-0 | ···················· | ···················· | ···················· | ···················· | ···················· | ···················· | |
| I | 1382 | hCoV-19/USA/TX-HMH-7038/2020\|EPI_ISL_546456\|2020-0 | ···················· | ·············A······ | ···················· | ···················· | ··············N····· | ···················· | |
| I | 1384 | hCoV-19/USA/TX-HMH-7050/2020\|EPI_ISL_546463\|2020-0 | ···················· | ···················· | ···················· | ···················· | NN············N····· | NN·N················ | 1 |
| I | 1385 | hCoV-19/USA/TX-HMH-7051/2020\|EPI_ISL_546464\|2020-0 | ···················· | ·············A······ | ···················· | ···················· | ···················· | ···················· | |
| I | 1386 | hCoV-19/USA/TX-HMH-7052/2020\|EPI_ISL_546465\|2020-0 | ···················· | ···················· | ···················· | ···················· | ···················· | ···················· | |
| I | 1387 | hCoV-19/USA/TX-HMH-7053/2020\|EPI_ISL_546466\|2020-0 | ···················· | ···················· | ···················· | ···················· | ···················· | ···················· | |
| I | 1388 | hCoV-19/USA/TX-HMH-7056/2020\|EPI_ISL_546468\|2020-0 | ···················· | ···················· | ···················· | ···················· | ···················· | ···················· | |
| I | 1389 | hCoV-19/USA/TX-HMH-7057/2020\|EPI_ISL_546469\|2020-0 | ···················· | ···················· | ···················· | ···················· | ···················· | ···················· | |
| I | 1392 | hCoV-19/USA/TX-HMH-7065/2020\|EPI_ISL_546475\|2020-0 | ···················· | ···················· | ···················· | ···················· | ···················· | ···················· | |
| I | 1393 | hCoV-19/USA/TX-HMH-7066/2020\|EPI_ISL_546476\|2020-0 | ···················· | ···················· | ···················· | ···················· | ···················· | ···················· | |
| I | 1394 | hCoV-19/USA/TX-HMH-7068/2020\|EPI_ISL_546477\|2020-0 | ···················· | ···················· | ···················· | ···················· | ···················· | ···················· | |
| I | 1395 | hCoV-19/USA/TX-HMH-7069/2020\|EPI_ISL_546478\|2020-0 | ···················· | ···················· | ···················· | ···················· | ··············N····· | NN·····N············ | 1 |
| I | 1396 | hCoV-19/USA/TX-HMH-7070/2020\|EPI_ISL_546479\|2020-0 | ···················· | ···················· | ···················· | ···················· | ···················· | ···················· | |
| I | 1397 | hCoV-19/USA/TX-HMH-7071/2020\|EPI_ISL_546480\|2020-0 | ···················· | ···················· | ···················· | ···················· | ···················· | ···················· | |
| I | 1398 | hCoV-19/USA/TX-HMH-7073/2020\|EPI_ISL_546482\|2020-0 | ···················· | ·············A······ | ···················· | ···················· | ···················· | ···················· | |
| I | 1399 | hCoV-19/USA/TX-HMH-7074/2020\|EPI_ISL_546483\|2020-0 | ···················· | ···················· | ···················· | ···················· | ··············N····· | N··················· | |

| | | | | | | | | | |
|---|---|---|---|---|---|---|---|---|---|
| I | 1400 | hCoV-19/USA/TX-HMH-7075/2020\|EPI_ISL_546484\|2020-0 | ···················· | ···················· | ···················· | ···················· | ···················· | ···················· | |
| I | 1401 | hCoV-19/USA/TX-HMH-7078/2020\|EPI_ISL_546486\|2020-0 | ···················· | ···················· | ···················· | ···················· | ···················· | ···················· | |
| I | 1402 | hCoV-19/USA/TX-HMH-7080/2020\|EPI_ISL_546487\|2020-0 | ···················· | ···················· | ···················· | ···················· | ···················· | ···················· | |
| I | 1403 | hCoV-19/USA/TX-HMH-7081/2020\|EPI_ISL_546488\|2020-0 | ···················· | ···················· | ···················· | ···················· | ···················· | ···················· | |
| I | 1404 | hCoV-19/USA/TX-HMH-7082/2020\|EPI_ISL_546489\|2020-0 | ···················· | ···················· | ···················· | ···················· | ···················· | ···················· | |
| I | 1405 | hCoV-19/USA/TX-HMH-7083/2020\|EPI_ISL_546490\|2020-0 | ···················· | ···················· | ···················· | ···················· | ·············NN····· | ······NN············ | |
| I | 1407 | hCoV-19/USA/TX-HMH-7089/2020\|EPI_ISL_546494\|2020-0 | ···················· | ···················· | ···················· | ···················· | ···················· | ···················· | |
| I | 1408 | hCoV-19/USA/TX-HMH-709/2020\|EPI_ISL_546495\|2020-04 | ···················· | ···················· | ···················· | ···················· | ···················· | ···················· | |
| I | 1409 | hCoV-19/USA/TX-HMH-7093/2020\|EPI_ISL_546498\|2020-0 | ···················· | ···················· | ···················· | ···················· | ···················· | ···················· | |
| I | 1410 | hCoV-19/USA/TX-HMH-7095/2020\|EPI_ISL_546499\|2020-0 | ···················· | ···················· | ···················· | ···················· | ·············N······ | ···················· | |
| I | 1411 | hCoV-19/USA/TX-HMH-5794/2020\|EPI_ISL_547020\|2020-0 | ···················· | ···················· | ···················· | ···················· | ···················· | ···················· | |
| I | 1412 | hCoV-19/USA/TX-HMH-5795/2020\|EPI_ISL_547021\|2020-0 | ···················· | ···················· | ···················· | ···················· | ···················· | ···················· | |
| I | 1413 | hCoV-19/USA/TX-HMH-5798/2020\|EPI_ISL_547022\|2020-0 | ···················· | ···················· | ···················· | ···················· | ···················· | ···················· | |
| I | 1416 | hCoV-19/USA/TX-HMH-5805/2020\|EPI_ISL_547027\|2020-0 | ···················· | ···················· | ···················· | ···················· | ···················· | ···················· | |
| I | 1417 | hCoV-19/USA/TX-HMH-5806/2020\|EPI_ISL_547028\|2020-0 | ···················· | ···················· | ···················· | ···················· | ···················· | ···················· | |
| I | 1418 | hCoV-19/USA/TX-HMH-5807/2020\|EPI_ISL_547029\|2020-0 | ···················· | ···················· | ···················· | ···················· | ···················· | ···················· | |
| I | 1419 | hCoV-19/USA/TX-HMH-5809/2020\|EPI_ISL_547031\|2020-0 | ···················· | ·A·················· | ···················· | ···················· | ···················· | ···················· | |
| I | 1420 | hCoV-19/USA/TX-HMH-5812/2020\|EPI_ISL_547034\|2020-0 | ···················· | ···················· | ···················· | ···················· | ···················· | ···················· | |
| I | 1421 | hCoV-19/USA/TX-HMH-5813/2020\|EPI_ISL_547035\|2020-0 | ···················· | ···················· | ···················· | ···················· | ·············N······ | ·······N············ | |
| I | 1423 | hCoV-19/USA/TX-HMH-5815/2020\|EPI_ISL_547037\|2020-0 | ···················· | ···················· | ···················· | ···················· | ···················· | ···················· | |
| I | 1424 | hCoV-19/USA/TX-HMH-5819/2020\|EPI_ISL_547040\|2020-0 | ···················· | ···················· | ···················· | ···················· | ···················· | ···················· | |
| I | 1429 | hCoV-19/USA/TX-HMH-5830/2020\|EPI_ISL_547046\|2020-0 | ···················· | ···················· | ···················· | ···················· | ···················· | ···················· | |
| I | 1431 | hCoV-19/USA/TX-HMH-5833/2020\|EPI_ISL_547048\|2020-0 | ···················· | ···················· | ···················· | ···················· | ···················· | ···················· | |
| I | 1436 | hCoV-19/USA/TX-HMH-5842/2020\|EPI_ISL_547056\|2020-0 | ···················· | ···················· | ···················· | ···················· | ···················· | ···················· | |
| I | 1439 | hCoV-19/USA/TX-HMH-5851/2020\|EPI_ISL_547062\|2020-0 | ···················· | ···················· | ···················· | ···················· | ···················· | ···················· | |
| I | 1444 | hCoV-19/USA/TX-HMH-586/2020\|EPI_ISL_547069\|2020-04 | ···················· | ···················· | ···················· | ···················· | ···················· | ···················· | |
| I | 1445 | hCoV-19/USA/TX-HMH-5862/2020\|EPI_ISL_547071\|2020-0 | ···················· | ············A······· | ···················· | ···················· | ···················· | ···················· | |
| I | 1446 | hCoV-19/USA/TX-HMH-5863/2020\|EPI_ISL_547072\|2020-0 | ···················· | ···················· | ···················· | ···················· | ·············N······ | NN·················· | |
| I | 1448 | hCoV-19/USA/TX-HMH-5866/2020\|EPI_ISL_547074\|2020-0 | ···················· | ···················· | ···················· | ···················· | ···················· | ···················· | |
| I | 1449 | hCoV-19/USA/TX-HMH-5870/2020\|EPI_ISL_547076\|2020-0 | ···················· | ···················· | ···················· | ···················· | ···················· | ···················· | |
| I | 1450 | hCoV-19/USA/TX-HMH-5873/2020\|EPI_ISL_547077\|2020-0 | ···················· | ············A······· | ···················· | ···················· | ···················· | ···················· | |
| I | 1451 | hCoV-19/USA/TX-HMH-5876/2020\|EPI_ISL_547079\|2020-0 | ···················· | ···················· | ···················· | ···················· | ···················· | ···················· | |
| I | 1452 | hCoV-19/USA/IA-GMF-18345/2020\|EPI_ISL_547758\|2020- | ···················· | ···················· | ···················· | ···················· | ···················· | ···················· | |
| I | 1453 | hCoV-19/USA/WI-GMF-18405/2020\|EPI_ISL_547759\|2020- | ···················· | ···················· | ···················· | ···················· | ···················· | ··············A····· | |
| I | 1454 | hCoV-19/USA/WI-GMF-18456/2020\|EPI_ISL_547760\|2020- | ···················· | ···················· | ···················· | ···················· | ···················· | ···················· | |
| I | 1455 | hCoV-19/USA/WI-GMF-18461/2020\|EPI_ISL_547761\|2020- | ···················· | ···················· | ···················· | ···················· | ···················· | ···················· | |
| I | 1456 | hCoV-19/USA/WI-GMF-18466/2020\|EPI_ISL_547762\|2020- | ···················· | ···················· | ···················· | ···················· | ···················· | ···················· | |
| I | 1457 | hCoV-19/USA/WI-GMF-18506/2020\|EPI_ISL_547763\|2020- | ···················· | ···················· | ···················· | ···················· | ···················· | ···················· | |
| I | 1458 | hCoV-19/USA/IA-GMF-18509/2020\|EPI_ISL_547764\|2020- | ···················· | ···················· | ···················· | ···················· | ···················· | ···················· | |
| I | 1459 | hCoV-19/USA/WI-GMF-18528/2020\|EPI_ISL_547765\|2020- | ···················· | ···················· | ···················· | ···················· | ···················· | ···················· | |
| I | 1460 | hCoV-19/USA/WI-GMF-18553/2020\|EPI_ISL_547766\|2020- | ···················· | ···················· | ···················· | ···················· | ···················· | ···················· | |
| I | 1461 | hCoV-19/USA/WI-GMF-18563/2020\|EPI_ISL_547767\|2020- | ···················· | ···················· | ···················· | ···················· | ···················· | ···················· | |
| I | 1462 | hCoV-19/USA/WI-GMF-18574/2020\|EPI_ISL_547768\|2020- | ···················· | ···················· | ···················· | ···················· | ···················· | ···················· | |
| I | 1463 | hCoV-19/USA/WI-GMF-18577/2020\|EPI_ISL_547769\|2020- | ···················· | ···················· | ···················· | ···················· | ···················· | ···················· | |
| I | 1464 | hCoV-19/USA/WI-GMF-18594/2020\|EPI_ISL_547770\|2020- | ···················· | ···················· | ···················· | ···················· | ···················· | ···················· | |
| I | 1465 | hCoV-19/USA/WI-GMF-18609/2020\|EPI_ISL_547771\|2020- | ···················· | ···················· | ···················· | ···················· | ···················· | ··············A····· | |
| I | 1466 | hCoV-19/USA/WI-GMF-18658/2020\|EPI_ISL_547772\|2020- | ···················· | ···················· | ···················· | ···················· | ···················· | ···················· | |
| I | 1467 | hCoV-19/USA/WI-GMF-18660/2020\|EPI_ISL_547773\|2020- | ···················· | ···················· | ···················· | ···················· | ···················· | ···················· | |
| I | 1468 | hCoV-19/USA/IL-GMF-18664/2020\|EPI_ISL_547774\|2020- | ···················· | ···················· | ···················· | ···················· | ···················· | ···················· | |
| I | 1469 | hCoV-19/USA/WI-GMF-18675/2020\|EPI_ISL_547775\|2020- | ···················· | ···················· | ···················· | ···················· | ···················· | ···················· | |
| I | 1470 | hCoV-19/USA/WI-GMF-18719/2020\|EPI_ISL_547776\|2020- | ···················· | ···················· | ···················· | ···················· | ···················· | ···················· | |
| I | 1471 | hCoV-19/USA/WI-GMF-18723/2020\|EPI_ISL_547777\|2020- | ···················· | ···················· | ···················· | ···················· | ···················· | ···················· | |
| I | 1472 | hCoV-19/USA/WI-GMF-18724/2020\|EPI_ISL_547778\|2020- | ···················· | ···················· | ···················· | ···················· | ···················· | ···················· | |
| I | 1473 | hCoV-19/USA/WI-GMF-18731/2020\|EPI_ISL_547779\|2020- | ···················· | ···················· | ···················· | ···················· | ···················· | ···················· | |
| I | 1474 | hCoV-19/USA/WI-GMF-18733/2020\|EPI_ISL_547780\|2020- | ···················· | ···················· | ···················· | ···················· | ···················· | ···················· | |
| I | 1475 | hCoV-19/USA/WI-GMF-18735/2020\|EPI_ISL_547781\|2020- | ···················· | ···················· | ···················· | ···················· | ···················· | ···················· | |
| I | 1476 | hCoV-19/USA/WI-GMF-18785/2020\|EPI_ISL_547782\|2020- | ···················· | ···················· | ···················· | ···················· | ···················· | ···················· | |
| I | 1477 | hCoV-19/USA/MN-GMF-18787/2020\|EPI_ISL_547783\|2020- | ···················· | ···················· | ···················· | ···················· | ···················· | ···················· | |
| I | 1478 | hCoV-19/USA/WI-GMF-18789/2020\|EPI_ISL_547784\|2020- | ···················· | ···················· | ···················· | ···················· | ···················· | ···················· | |
| I | 1479 | hCoV-19/USA/WI-GMF-18791/2020\|EPI_ISL_547785\|2020- | ···················· | ···················· | ···················· | ···················· | ···················· | ···················· | |
| I | 1480 | hCoV-19/USA/IA-GMF-18813/2020\|EPI_ISL_547786\|2020- | ···················· | ···················· | ···················· | ···················· | ···················· | ···················· | |
| I | 1481 | hCoV-19/USA/IA-GMF-18906/2020\|EPI_ISL_547787\|2020- | ···················· | ···················· | ···················· | ···················· | ···················· | ···················· | |
| I | 1482 | hCoV-19/USA/IA-GMF-18913/2020\|EPI_ISL_547788\|2020- | ···················· | ···················· | ···················· | ···················· | ···················· | ···················· | |
| I | 1483 | hCoV-19/USA/WI-GMF-18957/2020\|EPI_ISL_547789\|2020- | ···················· | ···················· | ···················· | ···················· | ···················· | ···················· | |
| I | 1484 | hCoV-19/USA/WI-GMF-18990/2020\|EPI_ISL_547790\|2020- | ···················· | ···················· | ···················· | ···················· | ···················· | ··············A····· | |
| I | 1485 | hCoV-19/USA/MN-GMF-19171/2020\|EPI_ISL_547791\|2020- | ···················· | ···················· | ···················· | ···················· | ···················· | ···················· | |
| I | 1486 | hCoV-19/USA/WI-GMF-19175/2020\|EPI_ISL_547792\|2020- | ···················· | ···················· | ···················· | ···················· | ···················· | ···················· | |
| I | 1487 | hCoV-19/USA/WI-GMF-19195/2020\|EPI_ISL_547793\|2020- | ···················· | ···················· | ···················· | ···················· | ···················· | ··············A····· | |
| I | 1488 | hCoV-19/USA/WI-GMF-19220/2020\|EPI_ISL_547794\|2020- | ···················· | ···················· | ···················· | ···················· | ···················· | ···················· | |
| I | 1489 | hCoV-19/USA/WI-GMF-19225/2020\|EPI_ISL_547795\|2020- | ···················· | ···················· | ···················· | ···················· | ···················· | ···················· | |
| I | 1490 | hCoV-19/USA/WI-GMF-19226/2020\|EPI_ISL_547796\|2020- | ···················· | ···················· | ···················· | ···················· | ···················· | ···················· | |
| I | 1491 | hCoV-19/USA/IA-GMF-19239/2020\|EPI_ISL_547797\|2020- | ···················· | ···················· | ···················· | ···················· | ···················· | ···················· | |
| I | 1492 | hCoV-19/USA/WI-GMF-19240/2020\|EPI_ISL_547798\|2020- | ···················· | ···················· | ···················· | ···················· | ···················· | ···················· | |
| I | 1493 | hCoV-19/Switzerland/GE-SNRCI-7799/2020\|EPI_ISL_548 | ···················· | ···················· | ···················· | ···················· | ···················· | ···················· | |
| I | 1494 | hCoV-19/Switzerland/GE-SNRCI-7808/2020\|EPI_ISL_548 | ···················· | ···················· | ···················· | ···················· | ···················· | ···················· | |
| I | 1495 | hCoV-19/Switzerland/GE-SNRCI-7836/2020\|EPI_ISL_548 | ···················· | ···················· | ···················· | ···················· | ···················· | ···················· | |
| I | 1496 | hCoV-19/Switzerland/GE-SNRCI-7891/2020\|EPI_ISL_548 | ···················· | ···················· | ···················· | ···················· | ···················· | ···················· | |
| I | 1497 | hCoV-19/Switzerland/GE-SNRCI-7895/2020\|EPI_ISL_548 | ···················· | ···················· | ···················· | ···················· | ···················· | ···················· | |
| I | 1498 | hCoV-19/Switzerland/GE-SNRCI-7899/2020\|EPI_ISL_548 | ···················· | ···················A | ···················· | ···················· | ···················· | ···················· | 1 |
| I | 1499 | hCoV-19/Switzerland/GE-SNRCI-7906/2020\|EPI_ISL_548 | ···················· | ···················· | ···················· | ···················· | ···················· | ···················· | |

| | | | | | | | | | |
|---|---|---|---|---|---|---|---|---|---|
| I | 1500 | hCoV-19/USA/CA-CZB-3642/2020\|EPI_ISL_548348\|2020-0 | ···················· | ···················· | ···················· | ···················· | ···················· | ···················· | |
| I | 1501 | hCoV-19/USA/CA-CZB-3638/2020\|EPI_ISL_548349\|2020-0 | ···················· | ···················· | ···················· | ···················· | ···················· | ···················· | |
| I | 1502 | hCoV-19/USA/CA-CZB-3632/2020\|EPI_ISL_548350\|2020-0 | ···················· | ···················· | ···················· | ···················· | ···················· | ···················· | |
| I | 1503 | hCoV-19/USA/CA-CZB-3629/2020\|EPI_ISL_548351\|2020-0 | ···················· | ···················· | ···················· | ···················· | ···················· | ···················· | |
| I | 1504 | hCoV-19/USA/CA-CZB-3647/2020\|EPI_ISL_548352\|2020-0 | ···················· | ···················· | ···················· | ···················· | ···················· | ···················· | |
| I | 1505 | hCoV-19/USA/CA-CZB-3645/2020\|EPI_ISL_548353\|2020-0 | ···················· | ···················· | ···················· | ···················· | ···················· | ···················· | |
| I | 1506 | hCoV-19/USA/CA-CZB-3636/2020\|EPI_ISL_548354\|2020-0 | ···················· | ···················· | ···················· | ···················· | ···················· | ···················· | |
| I | 1507 | hCoV-19/USA/CA-CZB-3641/2020\|EPI_ISL_548355\|2020-0 | ···················· | ···················· | ···················· | ···················· | ···················· | ···················· | |
| I | 1508 | hCoV-19/USA/CA-CZB-3637/2020\|EPI_ISL_548356\|2020-0 | ···················· | ···················· | ···················· | ···················· | ···················· | ···················· | |
| I | 1509 | hCoV-19/USA/CA-CZB-3631/2020\|EPI_ISL_548357\|2020-0 | ···················· | ···················· | ···················· | ···················· | ···················· | ···················· | |
| I | 1510 | hCoV-19/USA/CA-CZB-3644/2020\|EPI_ISL_548358\|2020-0 | ···················· | ···················· | ···················· | ···················· | ···················· | ···················· | |
| I | 1511 | hCoV-19/USA/CA-CZB-3633/2020\|EPI_ISL_548359\|2020-0 | ···················· | ·A·················· | ···················· | ···················· | ···················· | ···················· | |
| I | 1512 | hCoV-19/USA/CA-CZB-3625/2020\|EPI_ISL_548360\|2020-0 | ···················· | ·A·················· | ···················· | ···················· | ···················· | ···················· | |
| I | 1513 | hCoV-19/USA/CA-CZB-3630/2020\|EPI_ISL_548361\|2020-0 | ···················· | ···················· | ···················· | ···················· | ···················· | ···················· | |
| I | 1514 | hCoV-19/USA/CA-CZB-3643/2020\|EPI_ISL_548362\|2020-0 | ···················· | ···················· | ···················· | ···················· | ···················· | ···················· | |
| I | 1515 | hCoV-19/USA/CA-CZB-3673/2020\|EPI_ISL_548363\|2020-0 | ···················· | ·A·················· | ···················· | ···················· | ···················· | ···················· | |
| I | 1516 | hCoV-19/USA/CA-CZB-3646/2020\|EPI_ISL_548364\|2020-0 | ···················· | ···················· | ···················· | ···················· | ···················· | ···················· | |
| I | 1517 | hCoV-19/USA/CA-CZB-3627/2020\|EPI_ISL_548365\|2020-0 | ···················· | ···················· | ···················· | ···················· | ···················· | ···················· | |
| I | 1518 | hCoV-19/USA/CA-CZB-3624/2020\|EPI_ISL_548366\|2020-0 | ···················· | ·A·················· | ···················· | ···················· | ···················· | ···················· | |
| I | 1519 | hCoV-19/USA/CA-CZB-3635/2020\|EPI_ISL_548367\|2020-0 | ···················· | ···················· | ···················· | ···················· | ···················· | ···················· | |
| I | 1520 | hCoV-19/USA/CA-CZB-3639/2020\|EPI_ISL_548368\|2020-0 | ···················· | ···················· | ···················· | ···················· | ···················· | ···················· | |
| I | 1521 | hCoV-19/USA/CA-CZB-3628/2020\|EPI_ISL_548369\|2020-0 | ···················· | ···················· | ···················· | ···················· | ···················· | ···················· | |
| I | 1522 | hCoV-19/USA/CA-CZB-3626/2020\|EPI_ISL_548370\|2020-0 | ···················· | ···················· | ···················· | ···················· | ···················· | ···················· | |
| I | 1523 | hCoV-19/USA/CA-CZB-3739/2020\|EPI_ISL_548373\|2020-0 | ···················· | ·A·················· | ···················· | ···················· | ···················· | ···················· | |
| I | 1524 | hCoV-19/USA/CA-CZB-3680/2020\|EPI_ISL_548374\|2020-0 | ···················· | ···················· | ···················· | ···················· | ···················· | ···················· | |
| I | 1525 | hCoV-19/USA/CA-CZB-3757/2020\|EPI_ISL_548375\|2020-0 | ···················· | ···················· | ···················· | ···················· | ···················· | ···················· | |
| I | 1526 | hCoV-19/USA/CA-CZB-3794/2020\|EPI_ISL_548376\|2020-0 | ···················· | ···················· | ···················· | ···················· | ···················· | ···················· | |
| I | 1527 | hCoV-19/USA/CA-CZB-3732/2020\|EPI_ISL_548377\|2020-0 | ···················· | ···················· | ···················· | ···················· | ···················· | ···················· | |
| I | 1528 | hCoV-19/USA/CA-CZB-3851/2020\|EPI_ISL_548378\|2020-0 | ···················· | ···················· | ···················· | ···················· | ···················· | ···················· | |
| I | 1529 | hCoV-19/USA/CA-CZB-3709/2020\|EPI_ISL_548379\|2020-0 | ···················· | ···················· | ···················· | ···················· | ···················N | ···················· | 1 |
| I | 1530 | hCoV-19/USA/CA-CZB-3748/2020\|EPI_ISL_548381\|2020-0 | ···················· | ···················· | ···················· | ···················· | ···················· | ···················· | |
| I | 1531 | hCoV-19/USA/CA-CZB-3688/2020\|EPI_ISL_548382\|2020-0 | ···················· | ·A·················· | ···················· | ···················· | ···················· | ···················· | |
| I | 1532 | hCoV-19/USA/CA-CZB-3725/2020\|EPI_ISL_548386\|2020-0 | ···················· | ·A·················· | ···················· | ···················· | ··K················· | ···················· | |
| I | 1533 | hCoV-19/USA/CA-CZB-3758/2020\|EPI_ISL_548387\|2020-0 | ···················· | ···················· | ···················· | ···················· | ···················· | ···················· | |
| I | 1536 | hCoV-19/USA/CA-CZB-3777/2020\|EPI_ISL_548398\|2020-0 | ···················· | ·A·················· | ···················· | ···················· | ···················· | ···················· | |
| I | 1537 | hCoV-19/USA/TX-HMH-5278/2020\|EPI_ISL_544647\|2020-0 | ···················· | ···················· | ···················· | ···················· | ···················· | ···················· | |
| I | 1539 | hCoV-19/USA/TX-HMH-5280/2020\|EPI_ISL_544649\|2020-0 | ···················· | ···················· | ···················· | ···················· | ···················· | ···················· | |
| I | 1540 | hCoV-19/USA/TX-HMH-529/2020\|EPI_ISL_544656\|2020-04 | ···················· | ···················· | ···················· | ···················· | ···················· | ···················· | |
| I | 1541 | hCoV-19/USA/TX-HMH-5292/2020\|EPI_ISL_544658\|2020-0 | ···················· | ···················· | ···················· | ···················· | ···················· | ···················· | |
| I | 1542 | hCoV-19/USA/TX-HMH-5296/2020\|EPI_ISL_544659\|2020-0 | ···················· | ···················· | ···················· | ···················· | ···················· | ···················· | |
| I | 1543 | hCoV-19/USA/TX-HMH-5297/2020\|EPI_ISL_544660\|2020-0 | ···················· | ·············A······ | ···················· | ···················· | ···················· | NN·····N············ | 1 |
| I | 1544 | hCoV-19/USA/TX-HMH-5298/2020\|EPI_ISL_544661\|2020-0 | ···················· | ···················· | ···················· | ···················· | ···················· | ···················· | |
| I | 1546 | hCoV-19/USA/TX-HMH-5303/2020\|EPI_ISL_544664\|2020-0 | ···················· | ···················· | ···················· | ···················· | ···················· | ···················· | |
| I | 1548 | hCoV-19/USA/TX-HMH-5305/2020\|EPI_ISL_544666\|2020-0 | ···················· | ···················· | ···················· | ···················· | ···················· | NN·················· | |
| I | 1554 | hCoV-19/USA/TX-HMH-5313/2020\|EPI_ISL_544673\|2020-0 | ···················· | ···················· | ···················· | ···················· | ···················· | ···················· | |
| I | 1555 | hCoV-19/USA/TX-HMH-5316/2020\|EPI_ISL_544675\|2020-0 | ···················· | ···················· | ···················· | ···················· | ··············NN···· | ·······N···N········ | |
| I | 1556 | hCoV-19/USA/TX-HMH-5317/2020\|EPI_ISL_544676\|2020-0 | ···················· | ···················· | ···················· | ···················· | ···················· | ···················· | |
| I | 1557 | hCoV-19/USA/TX-HMH-5318/2020\|EPI_ISL_544677\|2020-0 | ···················· | ···················· | ···················· | ···················· | ···················· | ···················· | |
| I | 1559 | hCoV-19/USA/TX-HMH-5323/2020\|EPI_ISL_544680\|2020-0 | ···················· | ··········A········· | ···················· | ···················· | ···················· | ···················· | |
| I | 1560 | hCoV-19/USA/TX-HMH-5324/2020\|EPI_ISL_544681\|2020-0 | ···················· | ···················· | ···················· | ···················· | ···················· | ···················· | |
| I | 1561 | hCoV-19/USA/TX-HMH-5325/2020\|EPI_ISL_544682\|2020-0 | ···················· | ···················· | ···················· | ···················· | ···················· | ···················· | |
| I | 1562 | hCoV-19/USA/TX-HMH-5326/2020\|EPI_ISL_544683\|2020-0 | ···················· | ···················· | ···················· | ···················· | ···················· | ···················· | |
| I | 1563 | hCoV-19/USA/TX-HMH-5327/2020\|EPI_ISL_544684\|2020-0 | ···················· | ···················· | ···················· | ···················· | ···················· | ···················· | |
| I | 1564 | hCoV-19/USA/TX-HMH-533/2020\|EPI_ISL_544687\|2020-04 | ···················· | ···················· | ···················· | ···················· | ···················· | ···················· | |
| I | 1566 | hCoV-19/USA/TX-HMH-5333/2020\|EPI_ISL_544690\|2020-0 | ···················· | ···················· | ···················· | ···················· | ···················· | ···················· | |
| I | 1570 | hCoV-19/USA/TX-HMH-5338/2020\|EPI_ISL_544694\|2020-0 | ···················· | ···················· | ···················· | ···················· | ···················· | ···················· | |
| I | 1571 | hCoV-19/USA/TX-HMH-5339/2020\|EPI_ISL_544695\|2020-0 | ···················· | ···················· | ···················· | ···················· | ···················· | ···················· | |
| I | 1573 | hCoV-19/USA/TX-HMH-5341/2020\|EPI_ISL_544697\|2020-0 | ···················· | ·············A······ | ···················· | ···················· | ···················· | ···················· | |
| I | 1574 | hCoV-19/USA/TX-HMH-5342/2020\|EPI_ISL_544698\|2020-0 | ···················· | ···················· | ···················· | ···················· | ···················· | ···················· | |
| I | 1576 | hCoV-19/USA/TX-HMH-5344/2020\|EPI_ISL_544700\|2020-0 | ···················· | ···················· | ···················· | ···················· | ···················· | ···················· | |
| I | 1578 | hCoV-19/USA/TX-HMH-5347/2020\|EPI_ISL_544702\|2020-0 | ···················· | ···················· | ···················· | ···················· | ···················· | ···················· | |
| I | 1579 | hCoV-19/USA/TX-HMH-5348/2020\|EPI_ISL_544703\|2020-0 | ···················· | ···················· | ···················· | ···················· | ···················· | ···················· | |
| I | 1580 | hCoV-19/USA/TX-HMH-535/2020\|EPI_ISL_544705\|2020-04 | ···················· | ···················· | ···················· | ···················· | ···················· | ···················· | |
| I | 1581 | hCoV-19/USA/TX-HMH-5350/2020\|EPI_ISL_544706\|2020-0 | ···················· | ·············A······ | ···················· | ···················· | ···················· | ···················· | |
| I | 1582 | hCoV-19/USA/TX-HMH-5351/2020\|EPI_ISL_544707\|2020-0 | ···················· | ·············A······ | ···················· | ···················· | ···················· | ···················· | |
| I | 1583 | hCoV-19/USA/TX-HMH-5352/2020\|EPI_ISL_544708\|2020-0 | ···················· | ···················· | ···················· | ···················· | ···················· | ···················· | |
| I | 1585 | hCoV-19/USA/TX-HMH-5356/2020\|EPI_ISL_544711\|2020-0 | ···················· | ···················· | ···················· | ···················· | ···················· | ···················· | |
| I | 1587 | hCoV-19/USA/TX-HMH-536/2020\|EPI_ISL_544713\|2020-04 | ···················· | ···················· | ···················· | ···················· | ···················· | ···················· | |
| I | 1588 | hCoV-19/USA/TX-HMH-5360/2020\|EPI_ISL_544714\|2020-0 | ···················· | ···················· | ···················· | ···················· | ···················· | ···················· | |
| I | 1589 | hCoV-19/USA/TX-HMH-5361/2020\|EPI_ISL_544715\|2020-0 | ···················· | ···················· | ···················· | ···················· | ···················· | ···················· | |
| I | 1590 | hCoV-19/USA/TX-HMH-5362/2020\|EPI_ISL_544716\|2020-0 | ···················· | ···················· | ···················· | ···················· | ···················· | ···················· | |
| I | 1594 | hCoV-19/USA/TX-HMH-5366/2020\|EPI_ISL_544720\|2020-0 | ···················· | ·A·················· | ···················· | ···················· | ···················· | ···················· | |
| I | 1595 | hCoV-19/USA/TX-HMH-5367/2020\|EPI_ISL_544721\|2020-0 | ···················· | ·············A······ | ···················· | ···················· | ···················· | ···················· | |
| I | 1596 | hCoV-19/USA/TX-HMH-5370/2020\|EPI_ISL_544722\|2020-0 | ···················· | ···················· | ···················· | ···················· | ···················· | ···················· | |
| I | 1597 | hCoV-19/USA/TX-HMH-5371/2020\|EPI_ISL_544723\|2020-0 | ···················· | ·A·················· | ···················· | ···················· | ···················· | ···················· | |
| I | 1598 | hCoV-19/USA/TX-HMH-5372/2020\|EPI_ISL_544724\|2020-0 | ···················· | ···················· | ···················· | ···················· | ···················· | ···················· | |
| I | 1599 | hCoV-19/USA/TX-HMH-5373/2020\|EPI_ISL_544725\|2020-0 | ···················· | ···················· | ···················· | ···················· | ···················· | ···················· | |
| I | 1600 | hCoV-19/USA/TX-HMH-5374/2020\|EPI_ISL_544726\|2020-0 | ···················· | ··········A········· | ···················· | ···················· | ···················· | ···················· | |
| I | 1601 | hCoV-19/USA/TX-HMH-5375/2020\|EPI_ISL_544727\|2020-0 | ···················· | ···················· | ···················· | ···················· | ···················· | ···················· | |
| I | 1603 | hCoV-19/USA/TX-HMH-5378/2020\|EPI_ISL_544730\|2020-0 | ···················· | ···················· | ···················· | ···················· | ···················· | ···················· | |

| | | | | | | | | |
|---|---|---|---|---|---|---|---|---|
| I | 1604 hCoV-19/USA/TX-HMH-5381/2020\|EPI_ISL_544733\|2020-0 | ···················· | ···················· | ···················· | ···················· | ···················· | ···················· | |
| I | 1605 hCoV-19/USA/TX-HMH-5382/2020\|EPI_ISL_544734\|2020-0 | ···················· | ···················· | ···················· | ···················· | ···················· | ···················· | |
| I | 1606 hCoV-19/USA/TX-HMH-5387/2020\|EPI_ISL_544737\|2020-0 | ···················· | ···················· | ···················· | ···················· | ···················· | ···················· | |
| I | 1607 hCoV-19/USA/TX-HMH-5388/2020\|EPI_ISL_544738\|2020-0 | ···················· | ···················· | ···················· | ···················· | ···················· | ···················· | |
| I | 1608 hCoV-19/USA/TX-HMH-5389/2020\|EPI_ISL_544739\|2020-0 | ···················· | ···················· | ···················· | ···················· | ··············NNN··· | ········N··········· | 1 |
| I | 1609 hCoV-19/USA/TX-HMH-5393/2020\|EPI_ISL_544741\|2020-0 | ···················· | ····T··············· | ···················· | ···················· | ···················· | ···················· | |
| I | 1610 hCoV-19/USA/TX-HMH-5396/2020\|EPI_ISL_544742\|2020-0 | ···················· | ·············A······ | ···················· | ···················· | ···················· | ···················· | |
| I | 1614 hCoV-19/USA/TX-HMH-5403/2020\|EPI_ISL_544748\|2020-0 | ···················· | ·A·················· | ···················· | ···················· | ···················· | ···················· | |
| I | 1616 hCoV-19/USA/TX-HMH-5412/2020\|EPI_ISL_544756\|2020-0 | ···················· | ···················· | ···················· | ···················· | ···················· | ···················· | |
| I | 1618 hCoV-19/USA/TX-HMH-5415/2020\|EPI_ISL_544759\|2020-0 | ···················· | ···················· | ···················· | ···················· | ···················· | ···················· | |
| I | 1619 hCoV-19/USA/TX-HMH-5416/2020\|EPI_ISL_544760\|2020-0 | ···················· | ···················· | ···················· | ···················· | ···················· | ···················· | |
| I | 1620 hCoV-19/USA/TX-HMH-5418/2020\|EPI_ISL_544762\|2020-0 | ···················· | ···················· | ···················· | ···················· | ···················· | ···················· | |
| I | 1622 hCoV-19/USA/TX-HMH-5420/2020\|EPI_ISL_544764\|2020-0 | ···················· | ···················· | ···················· | ···················· | ···················· | ···················· | |
| I | 1624 hCoV-19/USA/TX-HMH-2203/2020\|EPI_ISL_545180\|2020-0 | ···················· | ···················· | ···················· | ···················· | ···················· | ···················· | |
| I | 1625 hCoV-19/USA/TX-HMH-2204/2020\|EPI_ISL_545181\|2020-0 | ···················· | ···················· | ···················· | ···················· | ···················· | ···················· | |
| I | 1626 hCoV-19/USA/TX-HMH-2208/2020\|EPI_ISL_545182\|2020-0 | ···················· | ···················· | ···················· | ···················· | ···················· | ···················· | |
| I | 1627 hCoV-19/USA/TX-HMH-2209/2020\|EPI_ISL_545183\|2020-0 | ···················· | ··············C····· | ···················· | ···················· | ···················· | ···················· | |
| I | 1628 hCoV-19/USA/TX-HMH-2211/2020\|EPI_ISL_545184\|2020-0 | ···················· | ···················· | ···················· | ···················· | ···················· | ···················· | |
| I | 1629 hCoV-19/USA/TX-HMH-2212/2020\|EPI_ISL_545185\|2020-0 | ···················· | ···················· | ···················· | ···················· | ···················· | ···················· | |
| I | 1630 hCoV-19/USA/TX-HMH-2213/2020\|EPI_ISL_545186\|2020-0 | ···················· | ···················· | ···················· | ···················· | ···················· | ···················· | |
| I | 1631 hCoV-19/USA/TX-HMH-2217/2020\|EPI_ISL_545187\|2020-0 | ···················· | ···················· | ···················· | ···················· | ··············N····· | ···A················ | |
| I | 1632 hCoV-19/USA/TX-HMH-2223/2020\|EPI_ISL_545188\|2020-0 | ···················· | ···················· | ···················· | ···················· | ···················· | ···················· | |
| I | 1633 hCoV-19/USA/TX-HMH-2224/2020\|EPI_ISL_545189\|2020-0 | ···················· | ···················· | ···················· | ···················· | ···················· | ···················· | |
| I | 1634 hCoV-19/USA/TX-HMH-2231/2020\|EPI_ISL_545191\|2020-0 | ···················· | ···················· | ···················· | ···················· | ···················· | ···················· | |
| I | 1635 hCoV-19/USA/TX-HMH-2233/2020\|EPI_ISL_545193\|2020-0 | ···················· | ···················· | ···················· | ···················· | ···················· | ···················· | |
| I | 1636 hCoV-19/USA/TX-HMH-2234/2020\|EPI_ISL_545194\|2020-0 | ···················· | ···················· | ···················· | ···················· | ···················· | ···················· | |
| I | 1637 hCoV-19/USA/TX-HMH-2235/2020\|EPI_ISL_545195\|2020-0 | ···················· | ···················· | ···················· | ···················· | ···················· | ···················· | |
| I | 1638 hCoV-19/USA/TX-HMH-2236/2020\|EPI_ISL_545196\|2020-0 | ···················· | ···················· | ···················· | ···················· | ···················· | ···················· | |
| I | 1639 hCoV-19/USA/TX-HMH-2238/2020\|EPI_ISL_545198\|2020-0 | ···················· | ···················· | ···················· | ···················· | ···················· | ···················· | |
| I | 1640 hCoV-19/USA/TX-HMH-2242/2020\|EPI_ISL_545199\|2020-0 | ···················· | ···················· | ···················· | ···················· | ···················· | ···················· | |
| I | 1641 hCoV-19/USA/TX-HMH-6421/2020\|EPI_ISL_545987\|2020-0 | ···················· | ···················· | ···················· | ···················· | ···················· | ···················· | |
| I | 1642 hCoV-19/USA/TX-HMH-6425/2020\|EPI_ISL_545988\|2020-0 | ···················· | ···················· | ···················· | ···················· | ···················· | ···················· | |
| I | 1643 hCoV-19/USA/TX-HMH-6426/2020\|EPI_ISL_545989\|2020-0 | ···················· | ···················· | ···················· | ···················· | ···················· | ···················· | |
| I | 1644 hCoV-19/USA/TX-HMH-6431/2020\|EPI_ISL_545991\|2020-0 | ···················· | ·A·················· | ···················· | ···················· | ···················· | ···················· | |
| I | 1646 hCoV-19/USA/TX-HMH-6435/2020\|EPI_ISL_545994\|2020-0 | ···················· | ···················· | ···················· | ···················· | ···················· | ···················· | |
| I | 1647 hCoV-19/USA/TX-HMH-6438/2020\|EPI_ISL_545995\|2020-0 | ···················· | ···················· | ···················· | ···················· | ···················· | ···················· | |
| I | 1648 hCoV-19/USA/TX-HMH-6441/2020\|EPI_ISL_545998\|2020-0 | ···················· | ···················· | ···················· | ···················· | ···················· | ···················· | |
| I | 1649 hCoV-19/USA/TX-HMH-6442/2020\|EPI_ISL_545999\|2020-0 | ···················· | ···················· | ···················· | ···················· | ······T············· | ···················· | |
| I | 1650 hCoV-19/USA/TX-HMH-6875/2020\|EPI_ISL_546310\|2020-0 | ···················· | ···················· | ···················· | ···················· | ···················· | ···················· | |
| I | 1651 hCoV-19/USA/TX-HMH-6975/2020\|EPI_ISL_546400\|2020-0 | ···················· | ···················· | ···················· | ···················· | ···················· | ···················· | |
| I | 1653 hCoV-19/USA/TX-HMH-6982/2020\|EPI_ISL_546406\|2020-0 | ···················· | ·············A······ | ···················· | ···················· | ··············N····· | ···················· | |
| I | 1655 hCoV-19/USA/TX-HMH-6985/2020\|EPI_ISL_546409\|2020-0 | ···················· | ···················· | ···················· | ···················· | ···················· | ···N················ | |
| I | 1659 hCoV-19/USA/TX-HMH-6991/2020\|EPI_ISL_546414\|2020-0 | ···················· | ···················· | ···················· | ···················· | ···················· | ···················· | |
| I | 1662 hCoV-19/USA/TX-HMH-7002/2020\|EPI_ISL_546423\|2020-0 | ···················· | ···················· | ···················· | ···················· | ···················· | ···················· | |
| I | 1664 hCoV-19/USA/TX-HMH-7006/2020\|EPI_ISL_546425\|2020-0 | ···················· | ·············A······ | ···················· | ···················· | ···················· | NN·················· | |
| I | 1665 hCoV-19/USA/TX-HMH-7007/2020\|EPI_ISL_546426\|2020-0 | ···················· | ···················· | ···················· | ···················· | ···················· | ···················· | |
| I | 1666 hCoV-19/USA/WI-GMF-15214/2020\|EPI_ISL_547701\|2020- | ···················· | ···················· | ···················· | ···················· | ···················· | ···················· | |
| I | 1667 hCoV-19/USA/WI-GMF-15269/2020\|EPI_ISL_547702\|2020- | ···················· | ·············A······ | ···················· | ···················· | ···················· | ···················· | |
| I | 1668 hCoV-19/USA/MN-GMF-15330/2020\|EPI_ISL_547703\|2020- | ···················· | ·············A······ | ···················· | ···················· | ···················· | ···················· | |
| I | 1669 hCoV-19/USA/IA-GMF-15466/2020\|EPI_ISL_547704\|2020- | ···················· | ···················· | ···················· | ···················· | ···················· | ···················· | |
| I | 1670 hCoV-19/USA/MN-GMF-M00029/2020\|EPI_ISL_547705\|2020 | ···················· | ·············A······ | ···················· | ···················· | ···················· | ···················· | |
| I | 1671 hCoV-19/USA/WI-GMF-15609/2020\|EPI_ISL_547706\|2020- | ···················· | ···················· | ···················· | ···················· | ···················· | ···················· | |
| I | 1672 hCoV-19/USA/WI-GMF-15652/2020\|EPI_ISL_547708\|2020- | ···················· | ···················· | ···················· | ···················· | ···················· | ···················· | |
| I | 1673 hCoV-19/USA/WI-GMF-15673/2020\|EPI_ISL_547709\|2020- | ···················· | ·············A······ | ···················· | ···················· | ···················· | ···················· | |
| I | 1674 hCoV-19/USA/MN-GMF-15765/2020\|EPI_ISL_547710\|2020- | ···················· | ···················· | ···················· | ···················· | ···················· | ···················· | |
| I | 1675 hCoV-19/USA/IA-GMF-15767/2020\|EPI_ISL_547711\|2020- | ···················· | ·············A······ | ···················· | ···················· | ···················· | ···················· | |
| I | 1676 hCoV-19/USA/WI-GMF-15909/2020\|EPI_ISL_547712\|2020- | ···················· | ···················· | ···················· | ···················· | ···················· | ···················· | |
| I | 1677 hCoV-19/USA/MN-GMF-15923/2020\|EPI_ISL_547713\|2020- | ···················· | ·A·················· | ···················· | ···················· | ···················· | ···················· | |
| I | 1678 hCoV-19/USA/WI-GMF-15972/2020\|EPI_ISL_547714\|2020- | ···················· | ···················· | ········G··········· | ···················· | ···················· | ···················· | |
| I | 1679 hCoV-19/USA/WI-GMF-15985/2020\|EPI_ISL_547715\|2020- | ···················· | ···················· | ···················· | ···················· | ···················· | ···················· | |
| I | 1680 hCoV-19/USA/WI-GMF-16096/2020\|EPI_ISL_547717\|2020- | ···················· | ···················· | ···················· | ···················· | ···················· | ···················· | |
| I | 1681 hCoV-19/USA/WI-GMF-16189/2020\|EPI_ISL_547718\|2020- | ···················· | ···················· | ···················· | ···················· | ···················· | ···················· | |
| I | 1682 hCoV-19/USA/WI-GMF-16207/2020\|EPI_ISL_547719\|2020- | ···················· | ···················· | ···················· | ···················· | ···················· | ···················· | |
| I | 1683 hCoV-19/USA/WI-GMF-16214/2020\|EPI_ISL_547720\|2020- | ···················· | ···················· | ···················· | ···················· | ···················· | ···················· | |
| I | 1684 hCoV-19/USA/WI-GMF-16244/2020\|EPI_ISL_547721\|2020- | ···················· | ···················· | ···················· | ···················· | ···················· | ···················· | |
| I | 1685 hCoV-19/USA/IA-GMF-16252/2020\|EPI_ISL_547722\|2020- | ···················· | ···················· | ···················· | ···················· | ···················· | ···················· | |
| I | 1686 hCoV-19/USA/WI-GMF-16264/2020\|EPI_ISL_547723\|2020- | ···················· | ···················· | ···················· | ···················· | ···················· | ···················· | |
| I | 1687 hCoV-19/USA/IA-GMF-16267/2020\|EPI_ISL_547724\|2020- | ···················· | ···················· | ···················· | ···················· | ···················· | ···················· | |
| I | 1688 hCoV-19/USA/WI-GMF-16473/2020\|EPI_ISL_547725\|2020- | ···················· | ···················· | ···················· | ···················· | ···················· | ···················· | |
| I | 1689 hCoV-19/USA/IA-GMF-16489/2020\|EPI_ISL_547726\|2020- | ···················· | ···················· | ···················· | ···················· | ···················· | ···················· | |
| I | 1690 hCoV-19/USA/WI-GMF-16632/2020\|EPI_ISL_547727\|2020- | ···················· | ···················· | ···················· | ···················· | ···················· | ···················· | |
| I | 1691 hCoV-19/USA/MN-GMF-16735/2020\|EPI_ISL_547728\|2020- | ···················· | ·············A······ | ···················· | ···················· | ···················· | ···················· | |
| I | 1692 hCoV-19/USA/WI-GMF-16793/2020\|EPI_ISL_547729\|2020- | ···················· | ···················· | ···················· | ···················· | ···················· | ···················· | |
| I | 1693 hCoV-19/USA/WI-GMF-16843/2020\|EPI_ISL_547730\|2020- | ···················· | ···················· | ···················· | ···················· | ···················· | ···················· | |
| I | 1694 hCoV-19/USA/WI-GMF-16858/2020\|EPI_ISL_547731\|2020- | ···················· | ···················· | ···················· | ···················· | ···················· | ···················· | |
| I | 1695 hCoV-19/USA/WI-GMF-16891/2020\|EPI_ISL_547732\|2020- | ···················· | ···················· | ···················· | ···················· | ···················· | ···················· | |
| I | 1696 hCoV-19/USA/MN-GMF-16946/2020\|EPI_ISL_547733\|2020- | ···················· | ·············A······ | ···················· | ···················· | ···················· | ···················· | |
| I | 1697 hCoV-19/USA/WI-GMF-17018/2020\|EPI_ISL_547734\|2020- | ···················· | ···················· | ···················· | ···················· | ···················· | ···················· | |
| I | 1698 hCoV-19/USA/IA-GMF-17036/2020\|EPI_ISL_547735\|2020- | ···················· | ·A·················· | ···················· | ···················· | ···················· | ···················· | |
| I | 1699 hCoV-19/USA/WI-GMF-17327/2020\|EPI_ISL_547736\|2020- | ···················· | ···················· | ···················· | ···················· | ···················· | ···················· | |

I 1700 hCoV-19/USA/WI-GMF-17427/2020|EPI_ISL_547740|2020- ···················· ·A·················· ····················· ···················· ···················· ····················
I 1701 hCoV-19/USA/MN-GMF-17485/2020|EPI_ISL_547741|2020- ···················· ·············A······ ····················· ···················· ···················· ····················
I 1702 hCoV-19/USA/MN-GMF-17505/2020|EPI_ISL_547742|2020- ···················· ···················· ····················· ···················· ···················· ····················
I 1703 hCoV-19/USA/WI-GMF-17507/2020|EPI_ISL_547743|2020- ···················· ·A·················· ····················· ···················· ···················· ····················
I 1704 hCoV-19/USA/WI-GMF-17526/2020|EPI_ISL_547744|2020- ···················· ···················· ····················· ···················· ···················· ····················
I 1705 hCoV-19/USA/WI-GMF-17570/2020|EPI_ISL_547746|2020- ···················· ···················· ····················· ···················· ···················· ····················
I 1706 hCoV-19/USA/WI-GMF-17647/2020|EPI_ISL_547747|2020- ···················· ···················· ····················· ···················· ···················· ····················
I 1707 hCoV-19/USA/WI-GMF-17808/2020|EPI_ISL_547748|2020- ···················· ···················· ····················· ···················· ···················· ····················
I 1708 hCoV-19/USA/WI-GMF-17866/2020|EPI_ISL_547749|2020- ···················· ···················· ····················· ···················· ···················· ····················
I 1709 hCoV-19/USA/WI-GMF-17921/2020|EPI_ISL_547750|2020- ···················· ···················· ····················· ···················· ···················· ····················
I 1710 hCoV-19/USA/WI-GMF-18003/2020|EPI_ISL_547751|2020- ···················· ···················· ····················· ···················· ···················· ····················
I 1711 hCoV-19/USA/WI-GMF-18031/2020|EPI_ISL_547752|2020- ···················· ···················· ····················· ···················· ···················· ····················
I 1712 hCoV-19/USA/WI-GMF-18080/2020|EPI_ISL_547753|2020- ···················· ···················· ····················· ···················· ···················· ····················
I 1713 hCoV-19/USA/IA-GMF-18219/2020|EPI_ISL_547754|2020- ···················· ···················· ····················· ···················· ···················· ····················
I 1714 hCoV-19/USA/WI-GMF-18264/2020|EPI_ISL_547755|2020- ···················· ···················· ····················· ···················· ···················· ····················
I 1715 hCoV-19/USA/WI-GMF-18277/2020|EPI_ISL_547756|2020- ···················· ·············A······ ····················· ···················· ···················· ····················
I 1716 hCoV-19/USA/WI-GMF-18312/2020|EPI_ISL_547757|2020- ···················· ···················· ····················· ···················· ···················· ····················
I 1717 hCoV-19/Switzerland/GE-SNRCI-7918/2020|EPI_ISL_548 ···················· ···················· ····················· ···················· ···················· ····················
I 1718 hCoV-19/Switzerland/GE-SNRCI-7927/2020|EPI_ISL_548 ···················· ···················· ····················· ···················· ···················· ····················
I 1719 hCoV-19/Switzerland/GE-SNRCI-7960/2020|EPI_ISL_548 ···················· ···················· ····················· ···················· ···················· ····················
I 1720 hCoV-19/Switzerland/GE-SNRCI-7986/2020|EPI_ISL_548 ···················· ·A·················· ····················· ···················· ···················· ····················
I 1721 hCoV-19/Switzerland/GE-SNRCI-8073/2020|EPI_ISL_548 ···················· ···················· ····················· ···················· ···················· ····················
I 1722 hCoV-19/Switzerland/GE-SNRCI-8083/2020|EPI_ISL_548 ···················· ···················· ····················· ···················· ···················· ····················
I 1723 hCoV-19/Switzerland/GE-SNRCI-8099/2020|EPI_ISL_548 ···················· ···················· ····················· ···················· ···················· ····················
I 1724 hCoV-19/Switzerland/GE-SNRCI-8123/2020|EPI_ISL_548 ···················· ·A·················· ····················· ···················· ···················· ····················
I 1725 hCoV-19/Switzerland/GE-SNRCI-8187/2020|EPI_ISL_548 ···················· ···················· ····················· ···················· ···················· ····················
I 1726 hCoV-19/Switzerland/GE-SNRCI-8226/2020|EPI_ISL_548 ···················· ···················· ····················· ···················· ···················· ····················
I 1727 hCoV-19/Switzerland/GE-SNRCI-8290/2020|EPI_ISL_548 ···················· ···················· ····················· ···················· ···················· ····················
I 1728 hCoV-19/Switzerland/GE-SNRCI-8298/2020|EPI_ISL_548 ···················· ···················· ····················· ···················· ···················· ····················
I 1729 hCoV-19/Switzerland/GE-SNRCI-8301/2020|EPI_ISL_548 ···················· ···················· ····················· ···················· ···················· ····················
I 1730 hCoV-19/Switzerland/GE-SNRCI-8323/2020|EPI_ISL_548 ···················· ···················· ····················· ···················· ···················· ····················
I 1731 hCoV-19/Switzerland/GE-SNRCI-8378/2020|EPI_ISL_548 ···················· ···················· ····················· ···················· ···················· ····················
I 1732 hCoV-19/Switzerland/GE-SNRCI-8420/2020|EPI_ISL_548 ···················· ······T············· ····················· ···················· ···················· ····················
I 1733 hCoV-19/Switzerland/GE-SNRCI-8431/2020|EPI_ISL_548 ···················· ···················· ····················· ···················· ···················· ····················
I 1734 hCoV-19/Switzerland/GE-SNRCI-8451/2020|EPI_ISL_548 ···················· ···················· ····················· ···················· ···················· ····················
I 1735 hCoV-19/Switzerland/GE-SNRCI-8452/2020|EPI_ISL_548 ···················· ·A·················· ····················· ···················· ···················· ····················
I 1736 hCoV-19/Switzerland/GE-SNRCI-8508/2020|EPI_ISL_548 ···················· ···················· ····················· ···················· ···················· ····················
I 1737 hCoV-19/Switzerland/GE-SNRCI-8715/2020|EPI_ISL_548 ···················· ···················· ····················· ···················· ···················· ····················
I 1738 hCoV-19/Switzerland/GE-SNRCI-8721/2020|EPI_ISL_548 ···················· ···················· ····················· ···················· ···················· ····················
I 1739 hCoV-19/Switzerland/GE-SNRCI-8820/2020|EPI_ISL_548 ···················· ···················· ····················· ···················· ···················· ····················
I 1740 hCoV-19/Switzerland/GE-SNRCI-8927/2020|EPI_ISL_548 ···················· ···················· ····················· ···················· ···················· ····················
I 1741 hCoV-19/Switzerland/GE-SNRCI-8936/2020|EPI_ISL_548 ···················· ···················· ····················· ···················· ···················· ····················
I 1742 hCoV-19/Switzerland/GE-SNRCI-9034/2020|EPI_ISL_548 ···················· ·A·················· ····················· ···················· ···················· ····················
I 1743 hCoV-19/Switzerland/GE-SNRCI-9136/2020|EPI_ISL_548 ···················· ···················· ····················· ···················· ···················· ····················
I 1744 hCoV-19/Switzerland/GE-SNRCI-9173/2020|EPI_ISL_548 ···················· ·A·················· ····················· ···················· ···················· ····················
I 1745 hCoV-19/Switzerland/GE-SNRCI-9980/2020|EPI_ISL_548 ···················· ···················· ····················· ···················· ···················· ····················
I 1746 hCoV-19/Switzerland/VD-SNRCI-8615/2020|EPI_ISL_548 ···················· ···················· ····················· ···················· ···················· ····················
I 1747 hCoV-19/Switzerland/VS-SNRCI-5838/2020|EPI_ISL_548 ···················· ···················· ····················· ···················· ···················· ····················
I 1748 hCoV-19/USA/NY-ISMMS-MH001827N/2020|EPI_ISL_548243 ···················· ···················· ····················· ···················· ···················· ····················
I 1749 hCoV-19/USA/NY-ISMMS-MH001832N/2020|EPI_ISL_548244 ···················· ···················· ····················· ···················· ···················· ····················
I 1750 hCoV-19/Sweden/20-08157/2020|EPI_ISL_548245|2020-0 ···················· ···················· ····················· ···················· ···················· ····················
I 1751 hCoV-19/Sweden/20-08186/2020|EPI_ISL_548246|2020-0 ···················· ···················· ····················· ···················· ···················· ····················
I 1752 hCoV-19/Sweden/20-08331/2020|EPI_ISL_548247|2020-0 ···················· ···················· ····················· ···················· ···················· ····················
I 1753 hCoV-19/Sweden/20-08830/2020|EPI_ISL_548248|2020-0 ···················· ···················· ····················· ···················· ···················· ····················
I 1754 hCoV-19/Sweden/20-50786/2020|EPI_ISL_548249|2020-0 ···················· ···················· ····················· ···················· ···················· ····················
I 1755 hCoV-19/Sweden/20-51742/2020|EPI_ISL_548250|2020-0 ···················· ···················· ····················· ···················· ···················· ····················
I 1756 hCoV-19/Sweden/20-51744/2020|EPI_ISL_548251|2020-0 ···················· ···················· ····················· ···················· ···················· ····················
I 1757 hCoV-19/Sweden/20-52034/2020|EPI_ISL_548252|2020-0 ···················· ···················· ····················· ···················· ···················· ····················
I 1758 hCoV-19/Sweden/20-52051/2020|EPI_ISL_548253|2020-0 ···················· ···················· ····················· ···················· ···················· ····················
I 1759 hCoV-19/Sweden/20-52053/2020|EPI_ISL_548254|2020-0 ···················· ···················· ····················· ···················· ···················· ····················
I 1760 hCoV-19/Sweden/20-52313/2020|EPI_ISL_548255|2020-0 ···················· ···················· ····················· ···················· ···················· ····················
I 1761 hCoV-19/Sweden/20-52355/2020|EPI_ISL_548256|2020-0 ···················· ···················· ····················· ···················· ···················· ····················
I 1762 hCoV-19/Sweden/20-52643/2020|EPI_ISL_548257|2020-0 ···················· ···················· ····················· ···················· ···················· ····················
I 1763 hCoV-19/Sweden/20-52650/2020|EPI_ISL_548258|2020-0 ···················· ···················· ····················· ···················· ···················· ····················
I 1764 hCoV-19/USA/CA-CZB-3295/2020|EPI_ISL_548264|2020-0 ···················· ···················· ····················· ···················· ···················· ····················
I 1765 hCoV-19/USA/CA-CZB-3297/2020|EPI_ISL_548265|2020-0 ···················· ···················· ····················· ···················· ···················· ····················
I 1766 hCoV-19/USA/CA-CZB-3289/2020|EPI_ISL_548266|2020-0 ···················· ·A·················· ····················· ···················· ···················· ····················
I 1767 hCoV-19/USA/CA-CZB-3269/2020|EPI_ISL_548267|2020-0 ···················· ···················· ····················· ···················· ···················· ····················
I 1768 hCoV-19/USA/CA-CZB-3317/2020|EPI_ISL_548268|2020-0 ···················· ·A·················· ····················· ···················· ···················· ····················
I 1769 hCoV-19/USA/CA-CZB-3307/2020|EPI_ISL_548269|2020-0 ···················· ···················· ····················· ···················· ···················· ····················
I 1770 hCoV-19/USA/CA-CZB-3344/2020|EPI_ISL_548270|2020-0 ···················· ·A·················· ····················· ···················· ···················· ····················
I 1771 hCoV-19/USA/CA-CZB-3339/2020|EPI_ISL_548271|2020-0 ···················· ·A·················· ····················· ···················· ···················· ····················
I 1772 hCoV-19/USA/CA-CZB-3333/2020|EPI_ISL_548272|2020-0 ···················· ·A·················· ····················· ···················· ···················· ····················
I 1773 hCoV-19/USA/CA-CZB-3351/2020|EPI_ISL_548273|2020-0 ···················· ···················· ····················· ···················· ···················· ····················
I 1774 hCoV-19/USA/CA-CZB-3362/2020|EPI_ISL_548274|2020-0 ···················· ···················· ····················· ···················· ···················· ····················
I 1775 hCoV-19/USA/CA-CZB-3343/2020|EPI_ISL_548275|2020-0 ···················· ·A·················· ····················· ···················· ···················· ····················
I 1776 hCoV-19/USA/CA-CZB-3332/2020|EPI_ISL_548276|2020-0 ···················· ···················· ····················· ···················· ···················· ····················
I 1777 hCoV-19/USA/CA-CZB-3279/2020|EPI_ISL_548279|2020-0 ···················· ···················· ····················· ···················· ···················· ····················
I 1778 hCoV-19/USA/CA-CZB-3318/2020|EPI_ISL_548280|2020-0 ···················· ···················· ····················· ···················· ···················· ····················
I 1783 hCoV-19/USA/CA-CZB-3352/2020|EPI_ISL_548285|2020-0 ···················· ···················· ····················· ···················· ···················· ····················

I 1785 hCoV-19/USA/CA-CZB-3303/2020|EPI_ISL_548288|2020-0 ···················· ···················· ···················· ···················· ···················· ····················
I 1786 hCoV-19/USA/CA-CZB-3364/2020|EPI_ISL_548289|2020-0 ···················· ·A·················· ···················· ···················· ···················· ····················
I 1787 hCoV-19/USA/CA-CZB-3302/2020|EPI_ISL_548292|2020-0 ···················· ···················· ···················· ···················· ···················· ····················
I 1788 hCoV-19/USA/CA-CZB-3325/2020|EPI_ISL_548294|2020-0 ···················· ···················· ···················· ···················· ···················· ····················
I 1790 hCoV-19/USA/CA-CZB-3347/2020|EPI_ISL_548296|2020-0 ···················· ···················· ···················· ···················· ···················· ····················
I 1791 hCoV-19/USA/CA-CZB-3350/2020|EPI_ISL_548298|2020-0 ···················· ···················· ···················· ···················· ···················· ····················
I 1792 hCoV-19/USA/CA-CZB-3286/2020|EPI_ISL_548299|2020-0 ···················· ···················· ···················· ···················· ···················· ····················
I 1793 hCoV-19/USA/TX-HMH-5099/2020|EPI_ISL_544524|2020-0 ···················· ···················· ···················· ···················· ···················· ····················
I 1794 hCoV-19/USA/TX-HMH-5104/2020|EPI_ISL_544527|2020-0 ···················· ···················· ···················· ···················· ···················· ····················
I 1795 hCoV-19/USA/TX-HMH-5105/2020|EPI_ISL_544528|2020-0 ···················· ·············A······ ···················· ···················· ···················· ····················
I 1798 hCoV-19/USA/TX-HMH-511/2020|EPI_ISL_544532|2020-04 ···················· ···················· ···················· ···················· ···················· ····················
I 1799 hCoV-19/USA/TX-HMH-5112/2020|EPI_ISL_544533|2020-0 ···················· ···················· ···················· ···················· ···················· ····················
I 1800 hCoV-19/USA/TX-HMH-5115/2020|EPI_ISL_544534|2020-0 ···················· ···················· ···················· ···················· ···················· ····················
I 1803 hCoV-19/USA/TX-HMH-512/2020|EPI_ISL_544538|2020-04 ···················· ···················· ···················· ···················· ···················· ····················
I 1804 hCoV-19/USA/TX-HMH-5126/2020|EPI_ISL_544541|2020-0 ···················· ···················· ···················· ···················· ··············N····· ····················
I 1805 hCoV-19/USA/TX-HMH-5127/2020|EPI_ISL_544542|2020-0 ···················· ···················· ···················· ···················· ··············NN···· NN·····N············ 1
I 1806 hCoV-19/USA/TX-HMH-5128/2020|EPI_ISL_544543|2020-0 ···················· ·············A······ ···················· ···················· ···················· ····················
I 1807 hCoV-19/USA/TX-HMH-513/2020|EPI_ISL_544545|2020-04 ···················· ···················· ···················· ···················· ···················· ····················
I 1809 hCoV-19/USA/TX-HMH-5132/2020|EPI_ISL_544548|2020-0 ···················· ···················· ···················· ···················· ···················· ····················
I 1811 hCoV-19/USA/TX-HMH-5138/2020|EPI_ISL_544551|2020-0 ···················· ·············A······ ···················· ···················· ···················· ····················
I 1812 hCoV-19/USA/TX-HMH-5140/2020|EPI_ISL_544552|2020-0 ···················· ···················· ···················· ···················· ···················· ····················
I 1813 hCoV-19/USA/TX-HMH-5142/2020|EPI_ISL_544553|2020-0 ···················· ···················· ···················· ···················· ···················· ·······NN···········
I 1814 hCoV-19/USA/TX-HMH-5145/2020|EPI_ISL_544556|2020-0 ···················· ···················· ···················· ···················· ···················· ····················
I 1816 hCoV-19/USA/TX-HMH-5152/2020|EPI_ISL_544561|2020-0 ···················· ···················· ···················· ···················· ···················· ····················
I 1817 hCoV-19/USA/TX-HMH-5153/2020|EPI_ISL_544562|2020-0 ···················· ···················· ···················· ···················· ···················· ····················
I 1821 hCoV-19/Australia/NSW919/2020|EPI_ISL_545000|2020- ···················· ···················· ···················· ···················· ···················· ····················
I 1822 hCoV-19/Australia/NSW920/2020|EPI_ISL_545001|2020- ···················· ···················· ···················· ···················· ···················· ····················
I 1823 hCoV-19/Australia/NSW921/2020|EPI_ISL_545002|2020- ···················· ···················· ···················· ···················· ···················· ····················
I 1824 hCoV-19/Australia/NSW922/2020|EPI_ISL_545003|2020- ···················· ···················· ···················· ···················· ···················· ····················
I 1825 hCoV-19/Australia/NSW923/2020|EPI_ISL_545004|2020- ···················· ···················· ···················· ···················· ···················· ····················
I 1826 hCoV-19/Australia/NSW924/2020|EPI_ISL_545005|2020- ···················· ···················· ···················· ···················· ···················· ····················
I 1827 hCoV-19/Australia/NSW925/2020|EPI_ISL_545006|2020- ···················· ···················· ···················· ···················· ···················· ····················
I 1828 hCoV-19/Australia/NSW926/2020|EPI_ISL_545007|2020- ···················· ···················· ···················· ···················· ···················· ····················
I 1829 hCoV-19/Australia/NSW927/2020|EPI_ISL_545008|2020- ···················· ···················· ···················· ···················· ···················· ····················
I 1830 hCoV-19/Australia/NSW937/2020|EPI_ISL_545009|2020- ···················· ···················· ···················· ···················· ···················· ····················
I 1831 hCoV-19/Australia/NSW938/2020|EPI_ISL_545010|2020- ···················· ···················· ···················· ···················· ···················· ····················
I 1832 hCoV-19/Australia/NSW939/2020|EPI_ISL_545011|2020- ···················· ···················· ···················· ···················· ···················· ····················
I 1833 hCoV-19/Australia/NSW940/2020|EPI_ISL_545012|2020- ···················· ···················· ···················· ···················· ···················· ····················
I 1834 hCoV-19/Australia/NSW941/2020|EPI_ISL_545013|2020- ···················· ···················· ···················· ···················· ···················· ····················
I 1835 hCoV-19/Australia/NSW942/2020|EPI_ISL_545014|2020- ···················· ···················· ···················· ···················· ···················· ····················
I 1836 hCoV-19/Australia/NSW943/2020|EPI_ISL_545015|2020- ···················· ···················· ···················· ···················· ···················· ····················
I 1837 hCoV-19/Australia/NSW944/2020|EPI_ISL_545016|2020- ···················· ···················· ···················· ···················· ···················· ····················
I 1838 hCoV-19/Australia/NSW945/2020|EPI_ISL_545017|2020- ···················· ···················· ···················· ···················· ···················· ····················
I 1839 hCoV-19/Australia/NSW946/2020|EPI_ISL_545018|2020- ···················· ···················· ···················· ···················· ···················· ····················
I 1840 hCoV-19/Australia/NSW947/2020|EPI_ISL_545019|2020- ···················· ···················· ···················· ···················· ···················· ····················
I 1841 hCoV-19/Australia/NSW948/2020|EPI_ISL_545020|2020- ···················· ···················· ···················· ···················· ···················· ····················
I 1842 hCoV-19/Australia/NSW949/2020|EPI_ISL_545021|2020- ···················· ···················· ···················· ···················· ···················· ····················
I 1843 hCoV-19/Australia/NSW950/2020|EPI_ISL_545022|2020- ···················· ···················· ···················· ···················· ···················· ····················
I 1844 hCoV-19/Australia/NSW951/2020|EPI_ISL_545023|2020- ···················· ···················· ···················· ···················· ···················· ····················
I 1845 hCoV-19/Australia/NSW952/2020|EPI_ISL_545024|2020- ········T··········· ···················· ···················· ···················· ···················· ····················
I 1846 hCoV-19/Australia/NSW953/2020|EPI_ISL_545025|2020- ········T··········· ···················· ···················· ···················· ···················· ····················
I 1847 hCoV-19/Australia/NSW954/2020|EPI_ISL_545026|2020- ···················· ···················· ···················· ···················· ···················· ····················
I 1848 hCoV-19/Australia/NSW955/2020|EPI_ISL_545027|2020- ···················· ···················· ···················· ···················· ···················· ····················
I 1849 hCoV-19/Australia/NSW956/2020|EPI_ISL_545028|2020- ········T··········· ···················· ···················· ···················· ···················· ····················
I 1850 hCoV-19/Australia/NSW957/2020|EPI_ISL_545029|2020- ········T··········· ···················· ···················· ···················· ···················· ····················
I 1851 hCoV-19/Australia/NSW958/2020|EPI_ISL_545030|2020- ···················· ···················· ···················· ···················· ···················· ····················
I 1852 hCoV-19/Australia/NSW959/2020|EPI_ISL_545031|2020- ···················· ···················· ···················· ···················· ···················· ····················
I 1853 hCoV-19/Australia/NSW960/2020|EPI_ISL_545032|2020- ········T··········· ···················· ···················· ···················· ···················· ····················
I 1854 hCoV-19/Australia/NSW961/2020|EPI_ISL_545033|2020- ········T··········· ···················· ···················· ···················· ···················· ····················
I 1855 hCoV-19/Australia/NSW962/2020|EPI_ISL_545034|2020- ···················· ···················· ···················· ···················· ···················· ····················
I 1856 hCoV-19/Australia/NSW963/2020|EPI_ISL_545035|2020- ···················· ···················· ···················· ···················· ···················· ····················
I 1857 hCoV-19/Australia/NSW964/2020|EPI_ISL_545036|2020- ···················· ···················· ···················· ···················· ···················· ····················
I 1858 hCoV-19/Australia/NSW965/2020|EPI_ISL_545037|2020- ···················· ···················· ···················· ···················· ···················· ····················
I 1859 hCoV-19/Australia/NSW966/2020|EPI_ISL_545038|2020- ········T··········· ···················· ···················· ···················· ···················· ····················
I 1860 hCoV-19/Australia/NSW967/2020|EPI_ISL_545039|2020- ········T··········· ···················· ···················· ···················· ···················· ····················
I 1861 hCoV-19/Australia/NSW968/2020|EPI_ISL_545040|2020- ···················· ···················· ···················· ···················· ···················· ····················
I 1862 hCoV-19/Australia/NSW969/2020|EPI_ISL_545041|2020- ········T··········· ···················· ···················· ···················· ···················· ····················
I 1863 hCoV-19/Australia/NSW970/2020|EPI_ISL_545042|2020- ···················· ···················· ···················· ···················· ···················· ····················
I 1864 hCoV-19/Australia/NSW971/2020|EPI_ISL_545043|2020- ········T··········· ···················· ···················· ···················· ···················· ····················
I 1865 hCoV-19/Australia/NSW972/2020|EPI_ISL_545044|2020- ········T··········· ···················· ···················· ···················· ···················· ····················
I 1866 hCoV-19/Australia/NSW973/2020|EPI_ISL_545045|2020- ···················· ···················· ···················· ···················· ···················· ····················
I 1867 hCoV-19/Australia/NSW974/2020|EPI_ISL_545046|2020- ········T··········· ···················· ···················· ···················· ···················· ····················
I 1868 hCoV-19/Australia/NSW975/2020|EPI_ISL_545047|2020- ········T··········· ···················· ···················· ···················· ···················· ····················
I 1869 hCoV-19/Australia/NSW976/2020|EPI_ISL_545048|2020- ···················· ···················· ···················· ···················· ···················· ····················
I 1870 hCoV-19/Australia/NSW977/2020|EPI_ISL_545049|2020- ········T··········· ···················· ···················· ···················· ···················· ····················
I 1871 hCoV-19/Australia/NSW978/2020|EPI_ISL_545050|2020- ···················· ···················· ···················· ···················· ···················· ····················
I 1872 hCoV-19/USA/TX-HMH-1983/2020|EPI_ISL_545051|2020-0 ···················· ······A············· ···················· ···················· ···················· ····················
I 1873 hCoV-19/USA/TX-HMH-1985/2020|EPI_ISL_545052|2020-0 ···················· ···················· ···················· ···················· ···················· ····················
I 1874 hCoV-19/USA/TX-HMH-1986/2020|EPI_ISL_545053|2020-0 ···················· ···················· ···················· ···················· ···················· ····················
I 1875 hCoV-19/USA/TX-HMH-1990/2020|EPI_ISL_545055|2020-0 ···················· ··············C····· ···················· ···················· ···················· ····················

| | | | | | | | |
|---|---|---|---|---|---|---|---|
| I | 1876 hCoV-19/USA/TX-HMH-1991/2020\|EPI_ISL_545056\|2020-0 | ···················· | ···················· | ···················· | ···················· | ···················· | ···················· |
| I | 1877 hCoV-19/USA/TX-HMH-1993/2020\|EPI_ISL_545057\|2020-0 | ···················· | ······A············· | ···················· | ···················· | ···················· | ···················· |
| I | 1878 hCoV-19/USA/TX-HMH-1996/2020\|EPI_ISL_545058\|2020-0 | ···················· | ···················· | ···················· | ···················· | ···················· | ···················· |
| I | 1879 hCoV-19/USA/TX-HMH-1999/2020\|EPI_ISL_545059\|2020-0 | ···················· | ···················· | ···················· | ···················· | ··············N····· | N··················· |
| I | 1880 hCoV-19/USA/TX-HMH-823/2020\|EPI_ISL_545855\|2020-04 | ···················· | ···················· | ···················· | ···················· | ···················· | ···················· |
| I | 1881 hCoV-19/USA/TX-HMH-824/2020\|EPI_ISL_545856\|2020-04 | ···················· | ···················· | ···················· | ···················· | ···················· | ···················· |
| I | 1882 hCoV-19/USA/TX-HMH-830/2020\|EPI_ISL_545857\|2020-04 | ···················· | ···················· | ···················· | ···················· | ···················· | ···················· |
| I | 1884 hCoV-19/USA/TX-HMH-836/2020\|EPI_ISL_545859\|2020-04 | ···················· | ···················· | ···················· | ···················· | ···················· | ···················· |
| I | 1885 hCoV-19/USA/TX-HMH-837/2020\|EPI_ISL_545860\|2020-04 | ···················· | ···················· | ···················· | ···················· | ···················· | ···················· |
| I | 1886 hCoV-19/USA/TX-HMH-838/2020\|EPI_ISL_545861\|2020-04 | ···················· | ···················· | ···················· | ···················· | ···················· | ···················· |
| I | 1889 hCoV-19/USA/TX-HMH-844/2020\|EPI_ISL_545865\|2020-04 | ···················· | ···················· | ···················· | ···················· | ···················· | ···················· |
| I | 1890 hCoV-19/USA/TX-HMH-846/2020\|EPI_ISL_545867\|2020-04 | ···················· | ···················· | ···················· | ···················· | ···················· | ···················· |
| I | 1891 hCoV-19/USA/TX-HMH-848/2020\|EPI_ISL_545868\|2020-04 | ···················· | ···················· | ···················· | ···················· | ···················· | ···················· |
| I | 1892 hCoV-19/USA/TX-HMH-849/2020\|EPI_ISL_545869\|2020-04 | ···················· | ···················· | ···················· | ···················· | ···················· | ···················· |
| I | 1893 hCoV-19/USA/TX-HMH-850/2020\|EPI_ISL_545870\|2020-04 | ···················· | ···················· | ···················· | ···················· | ···················· | ···················· |
| I | 1895 hCoV-19/USA/TX-HMH-853/2020\|EPI_ISL_545873\|2020-04 | ···················· | ···················· | ···················· | ···················· | ···················· | ···················· |
| I | 1896 hCoV-19/USA/TX-HMH-854/2020\|EPI_ISL_545874\|2020-04 | ···················· | ···················· | ···················· | ···················· | ···················· | ···················· |
| I | 1897 hCoV-19/USA/TX-HMH-855/2020\|EPI_ISL_545875\|2020-04 | ···················· | ···················· | ···················· | ···················· | ···················· | ···················· |
| I | 1898 hCoV-19/USA/TX-HMH-858/2020\|EPI_ISL_545876\|2020-04 | ···················· | ···················· | ···················· | ···················· | ···················· | ···················· |
| I | 1899 hCoV-19/USA/TX-HMH-860/2020\|EPI_ISL_545877\|2020-04 | ···················· | ···················· | ···················· | ···················· | ···················· | ···················· |
| I | 1900 hCoV-19/USA/TX-HMH-861/2020\|EPI_ISL_545878\|2020-04 | ···················· | ···················· | ···················· | ···················· | ···················· | ···················· |
| I | 1901 hCoV-19/USA/TX-HMH-862/2020\|EPI_ISL_545879\|2020-04 | ···················· | ···················· | ···················· | ···················· | ···················· | ···················· |
| I | 1902 hCoV-19/USA/TX-HMH-863/2020\|EPI_ISL_545880\|2020-04 | ···················· | ···················· | ···················· | ···················· | ···················· | ···················· |
| I | 1903 hCoV-19/USA/TX-HMH-864/2020\|EPI_ISL_545881\|2020-04 | ···················· | ···················· | ···················· | ···················· | ···················· | ···················· |
| I | 1904 hCoV-19/USA/TX-HMH-865/2020\|EPI_ISL_545882\|2020-04 | ···················· | ···················· | ···················· | ···················· | ···················· | ···················· |
| I | 1905 hCoV-19/USA/TX-HMH-866/2020\|EPI_ISL_545883\|2020-04 | ···················· | ···················· | ···················· | ···················· | ···················· | ···················· |
| I | 1906 hCoV-19/USA/TX-HMH-867/2020\|EPI_ISL_545884\|2020-04 | ···················· | ···················· | ···················· | ···················· | ···················· | ···················· |
| I | 1908 hCoV-19/USA/TX-HMH-869/2020\|EPI_ISL_545886\|2020-04 | ···················· | ···················· | ···················· | ···················· | ···················· | ···················· |
| I | 1909 hCoV-19/USA/TX-HMH-870/2020\|EPI_ISL_545887\|2020-04 | ···················· | ···················· | ···················· | ···················· | ···················· | ···················· |
| I | 1910 hCoV-19/USA/TX-HMH-871/2020\|EPI_ISL_545888\|2020-04 | ···················· | ···················· | ···················· | ···················· | ···················· | ···················· |
| I | 1911 hCoV-19/USA/TX-HMH-873/2020\|EPI_ISL_545890\|2020-04 | ···················· | ···················· | ···················· | ···················· | ···················· | ···················· |
| I | 1912 hCoV-19/USA/TX-HMH-874/2020\|EPI_ISL_545891\|2020-04 | ···················· | ···················· | ···················· | ···················· | ···················· | ···················· |
| I | 1913 hCoV-19/USA/TX-HMH-875/2020\|EPI_ISL_545892\|2020-04 | ···················· | ···················· | ···················· | ···················· | ···················· | ···················· |
| I | 1914 hCoV-19/USA/TX-HMH-876/2020\|EPI_ISL_545893\|2020-04 | ···················· | ···················· | ···················· | ···················· | ···················· | ···················· |
| I | 1915 hCoV-19/USA/TX-HMH-877/2020\|EPI_ISL_545894\|2020-04 | ···················· | ···················· | ···················· | ···················· | ···················· | ···················· |
| I | 1916 hCoV-19/USA/TX-HMH-881/2020\|EPI_ISL_545895\|2020-04 | ···················· | ···G················ | ···················· | ···················· | ···················· | ···················· |
| I | 1917 hCoV-19/USA/TX-HMH-884/2020\|EPI_ISL_545897\|2020-04 | ···················· | ···················· | ···················· | ···················· | ···················· | ···················· |
| I | 1918 hCoV-19/USA/TX-HMH-885/2020\|EPI_ISL_545898\|2020-04 | ···················· | ···················· | ···················· | ···················· | ···················· | ···················· |
| I | 1919 hCoV-19/USA/TX-HMH-6870/2020\|EPI_ISL_546306\|2020-0 | ···················· | ···················· | ···················· | ···················· | ··············N····· | NN·················· |
| I | 1920 hCoV-19/USA/TX-HMH-6872/2020\|EPI_ISL_546308\|2020-0 | ···················· | ···················· | ···················· | ···················· | ···················· | ···················· |
| I | 1922 hCoV-19/USA/TX-HMH-6876/2020\|EPI_ISL_546311\|2020-0 | ···················· | ···················· | ···················· | ···················· | ···················· | ···················· |
| I | 1923 hCoV-19/USA/TX-HMH-6877/2020\|EPI_ISL_546312\|2020-0 | ···················· | ···················· | ···················· | ···················· | ···················· | ···················· |
| I | 1924 hCoV-19/USA/TX-HMH-6878/2020\|EPI_ISL_546313\|2020-0 | ···················· | ·············A······ | ···················· | ···················· | ···················· | ···················· |
| I | 1925 hCoV-19/USA/TX-HMH-6879/2020\|EPI_ISL_546314\|2020-0 | ···················· | ···················· | ···················· | ···················· | ···················· | ···N················ |
| I | 1926 hCoV-19/USA/TX-HMH-688/2020\|EPI_ISL_546315\|2020-04 | ···················· | ···················· | ···················· | ···················· | ···················· | ···················· |
| I | 1927 hCoV-19/USA/TX-HMH-6881/2020\|EPI_ISL_546316\|2020-0 | ···················· | ·············A······ | ···················· | ···················· | ···················· | ···················· |
| I | 1930 hCoV-19/USA/TX-HMH-6884/2020\|EPI_ISL_546319\|2020-0 | ···················· | ···················· | ···················· | ···················· | ···················· | ···················· |
| I | 1931 hCoV-19/USA/TX-HMH-6885/2020\|EPI_ISL_546320\|2020-0 | ···················· | ···················· | ···················· | ···················· | ··············N····· | ···N···N············ |
| I | 1932 hCoV-19/USA/TX-HMH-6887/2020\|EPI_ISL_546322\|2020-0 | ···················· | ·············A······ | ···················· | ···················· | ···················· | ···················· |
| I | 1934 hCoV-19/USA/TX-HMH-689/2020\|EPI_ISL_546324\|2020-04 | ···················· | ···················· | ···················· | ···················· | ···················· | ···················· |
| I | 1935 hCoV-19/USA/TX-HMH-6892/2020\|EPI_ISL_546326\|2020-0 | ···················· | ···················· | ···················· | ···················· | ···················· | ···················· |
| I | 1937 hCoV-19/USA/TX-HMH-6894/2020\|EPI_ISL_546328\|2020-0 | ···················· | ···················· | ···················· | ···················· | ···················· | ···················· |
| I | 1940 hCoV-19/USA/TX-HMH-6898/2020\|EPI_ISL_546332\|2020-0 | ···················· | ···················· | ···················· | ···················· | ···················· | ···················· |
| I | 1941 hCoV-19/USA/TX-HMH-6899/2020\|EPI_ISL_546333\|2020-0 | ···················· | ·A·················· | ···················· | ···················· | ···················· | ···················· |
| I | 1942 hCoV-19/USA/TX-HMH-690/2020\|EPI_ISL_546334\|2020-04 | ···················· | ···················· | ···················· | ···················· | ··N················· | ···················· |
| I | 1943 hCoV-19/USA/TX-HMH-6900/2020\|EPI_ISL_546335\|2020-0 | ···················· | ···················· | ···················· | ···················· | ···················· | ···················· |
| I | 1944 hCoV-19/USA/TX-HMH-6901/2020\|EPI_ISL_546336\|2020-0 | ···················· | ···················· | ···················· | ···················· | ···················· | ···················· |
| I | 1945 hCoV-19/USA/TX-HMH-6902/2020\|EPI_ISL_546337\|2020-0 | ···················· | ·············A······ | ···················· | ···················· | ···················· | ···················· |
| I | 1949 hCoV-19/USA/TX-HMH-691/2020\|EPI_ISL_546342\|2020-04 | ···················· | ···················· | ···················· | ···················· | ···················· | ···················· |
| I | 1953 hCoV-19/USA/TX-HMH-692/2020\|EPI_ISL_546352\|2020-04 | ···················· | ···················· | ···················· | ···················· | ···················· | ···················· |
| I | 1959 hCoV-19/USA/TX-HMH-6931/2020\|EPI_ISL_546361\|2020-0 | ···················· | ·············A······ | ···················· | ···················· | ···················· | ···················· |
| I | 1961 hCoV-19/USA/TX-HMH-6934/2020\|EPI_ISL_546363\|2020-0 | ···················· | ···················· | ···················· | ···················· | ···················· | ·······N············ |
| I | 1962 hCoV-19/USA/TX-HMH-6937/2020\|EPI_ISL_546365\|2020-0 | ···················· | ···················· | ···················· | ···················· | ···················· | ···················· |
| I | 1964 hCoV-19/USA/TX-HMH-694/2020\|EPI_ISL_546368\|2020-04 | ···················· | ···················· | ···················· | ···················· | ···················· | ···················· |
| I | 1966 hCoV-19/USA/TX-HMH-6943/2020\|EPI_ISL_546372\|2020-0 | ···················· | ···················· | ···················· | ···················· | ···················· | ···················· |
| I | 1967 hCoV-19/USA/TX-HMH-6945/2020\|EPI_ISL_546373\|2020-0 | ···················· | ···················· | ···················· | ···················· | ···················· | ···················· |
| I | 1970 hCoV-19/USA/TX-HMH-6951/2020\|EPI_ISL_546379\|2020-0 | ···················· | ·············A······ | ···················· | ···················· | ···················· | ···················· |
| I | 1972 hCoV-19/USA/TX-HMH-6956/2020\|EPI_ISL_546384\|2020-0 | ···················· | ···················· | ···················· | ···················· | ···················· | ···N················ |
| I | 1973 hCoV-19/USA/TX-HMH-6958/2020\|EPI_ISL_546385\|2020-0 | ···················· | ···················· | ···················· | ···················· | ···················· | ···················· |
| I | 1974 hCoV-19/USA/TX-HMH-696/2020\|EPI_ISL_546386\|2020-04 | ···················· | ···················· | ···················· | ···················· | ···················· | ···················· |
| I | 1975 hCoV-19/USA/WI-GMF-11949/2020\|EPI_ISL_547637\|2020- | ···················· | ···················· | ···················· | ···················· | ···················· | ···················· |
| I | 1976 hCoV-19/USA/WI-GMF-12045/2020\|EPI_ISL_547638\|2020- | ···················· | ···················· | ···················· | ···················· | ···················· | ···················· |
| I | 1977 hCoV-19/USA/WI-GMF-12079/2020\|EPI_ISL_547639\|2020- | ···················· | ···················· | ···················· | ···················· | ···················· | ···················· |
| I | 1978 hCoV-19/USA/WI-GMF-12260/2020\|EPI_ISL_547640\|2020- | ···················· | ···················· | ···················· | ···················· | ···················· | ···················· |
| I | 1979 hCoV-19/USA/WI-GMF-12278/2020\|EPI_ISL_547641\|2020- | ···················· | ···················· | ···················· | ···················· | ···················· | ···················· |
| I | 1980 hCoV-19/USA/WI-GMF-12532/2020\|EPI_ISL_547642\|2020- | ···················· | ···················· | ···················· | ···················· | ···················· | ···················· |
| I | 1981 hCoV-19/USA/IA-GMF-12677/2020\|EPI_ISL_547643\|2020- | ···················· | ···················· | ···················· | ···················· | ···················· | ···················· |
| I | 1982 hCoV-19/USA/WI-GMF-12857/2020\|EPI_ISL_547644\|2020- | ···················· | ···················· | ···················· | ···················· | ···················· | ···················· |
| I | 1983 hCoV-19/USA/WI-GMF-12894/2020\|EPI_ISL_547645\|2020- | ···················· | ···················· | ···················· | ····N··············· | ···················· | ···················· |
| I | 1984 hCoV-19/USA/WI-GMF-12925/2020\|EPI_ISL_547646\|2020- | ···················· | ···················· | ···················· | ···················· | ···················· | ···················· |

| | | | | | | | | | |
|---|---|---|---|---|---|---|---|---|---|
| I | 1985 hCoV-19/USA/WI-GMF-12930/2020\|EPI_ISL_547647\|2020- | ···················· | ·A·················· | ····················· | ···················· | ···················· | ···················· | | |
| I | 1986 hCoV-19/USA/WI-GMF-12945/2020\|EPI_ISL_547648\|2020- | ···················· | ···················· | ····················· | ···················· | ···················· | ···················· | | |
| I | 1987 hCoV-19/USA/WI-GMF-13006/2020\|EPI_ISL_547649\|2020- | ···················· | ·A·················· | ····················· | ···················· | ···················· | ···················· | | |
| I | 1988 hCoV-19/USA/WI-GMF-13029/2020\|EPI_ISL_547650\|2020- | ···················· | ···················· | ····················· | ···················· | ···················· | ···················· | | |
| I | 1989 hCoV-19/USA/WI-GMF-13030/2020\|EPI_ISL_547651\|2020- | ···················· | ·A·················· | ····················· | ···················· | ···················· | ···················· | | |
| I | 1990 hCoV-19/USA/WI-GMF-13033/2020\|EPI_ISL_547652\|2020- | ···················· | ···················· | ····················· | ···················· | ···················· | ···················· | | |
| I | 1991 hCoV-19/USA/WI-GMF-13036/2020\|EPI_ISL_547653\|2020- | ···················· | ···················· | ····················· | ···················· | ···················· | ···················· | | |
| I | 1992 hCoV-19/USA/WI-GMF-13139/2020\|EPI_ISL_547654\|2020- | ···················· | ···················· | ····················· | ···················· | ···················· | ···················· | | |
| I | 1993 hCoV-19/USA/WI-GMF-13319/2020\|EPI_ISL_547655\|2020- | ···················· | ···················· | ····················· | ···················· | ···················· | ···················· | | |
| I | 1994 hCoV-19/USA/WI-GMF-13352/2020\|EPI_ISL_547656\|2020- | ···················· | ···················· | ····················· | ···················· | ···················· | ···················· | | |
| I | 1995 hCoV-19/USA/IA-GMF-13403/2020\|EPI_ISL_547657\|2020- | ···················· | ·············A······ | ····················· | ···················· | ···················· | ···················· | | |
| I | 1996 hCoV-19/USA/WI-GMF-13783/2020\|EPI_ISL_547658\|2020- | ···················· | ···················· | ····················· | ···················· | ···················· | ···················· | | |
| I | 1997 hCoV-19/USA/MN-GMF-13803/2020\|EPI_ISL_547659\|2020- | ···················· | ·············A······ | ····················· | ···················· | ···················· | ···················· | | |
| I | 1998 hCoV-19/USA/WI-GMF-13831/2020\|EPI_ISL_547660\|2020- | ···················· | ···················· | ····················· | ···················· | ···················· | ···················· | | |
| I | 1999 hCoV-19/USA/MN-GMF-13847/2020\|EPI_ISL_547661\|2020- | ···················· | ···················· | ····················· | ···················· | ···················· | ···················· | | |
| I | 2000 hCoV-19/USA/IA-GMF-13859/2020\|EPI_ISL_547662\|2020- | ···················· | ···················· | ····················· | ···················· | ···················· | ···················· | | |
| I | 2001 hCoV-19/USA/WI-GMF-13999/2020\|EPI_ISL_547664\|2020- | ···················· | ···················· | ····················· | ···················· | ···················· | ···················· | | |
| I | 2002 hCoV-19/USA/IA-GMF-14006/2020\|EPI_ISL_547665\|2020- | ···················· | ···················· | ····················· | ···················· | ···················· | ···················· | | |
| I | 2003 hCoV-19/USA/WI-GMF-M00015/2020\|EPI_ISL_547666\|2020 | ···················· | ···················· | ····················· | ···················· | ···················· | ···················· | | |
| I | 2004 hCoV-19/USA/WI-GMF-M00016/2020\|EPI_ISL_547667\|2020 | ···················· | ···················· | ····················· | ···················· | ···················· | ···················· | | |
| I | 2005 hCoV-19/USA/WI-GMF-M00017/2020\|EPI_ISL_547668\|2020 | ···················· | ···················· | ····················· | ···················· | ···················· | ···················· | | |
| I | 2006 hCoV-19/USA/WI-GMF-M00018/2020\|EPI_ISL_547669\|2020 | ···················· | ···················· | ····················· | ···················· | ···················· | ···················· | | |
| I | 2007 hCoV-19/USA/WI-GMF-M00019/2020\|EPI_ISL_547670\|2020 | ···················· | ···················· | ····················· | ···················· | ···················· | ···················· | | |
| I | 2008 hCoV-19/USA/WI-GMF-M00020/2020\|EPI_ISL_547671\|2020 | ···················· | ···················· | ····················· | ···················· | ···················· | ···················· | | |
| I | 2009 hCoV-19/USA/WI-GMF-M00021/2020\|EPI_ISL_547672\|2020 | ···················· | ···················· | ····················· | ···················· | ···················· | ···················· | | |
| I | 2010 hCoV-19/USA/WI-GMF-M00022/2020\|EPI_ISL_547673\|2020 | ···················· | ···················· | ····················· | ···················· | ···················· | ···················· | | |
| I | 2011 hCoV-19/USA/WI-GMF-M00023/2020\|EPI_ISL_547674\|2020 | ···················· | ···················· | ····················· | ···················· | ···················· | ···················· | | |
| I | 2012 hCoV-19/USA/WI-GMF-M00024/2020\|EPI_ISL_547675\|2020 | ···················· | ···················· | ····················· | ···················· | ···················· | ···················· | | |
| I | 2013 hCoV-19/USA/WI-GMF-M00026/2020\|EPI_ISL_547676\|2020 | ···················· | ···················· | ····················· | ···················· | ···················· | ···················· | | |
| I | 2014 hCoV-19/USA/WI-GMF-14101/2020\|EPI_ISL_547678\|2020- | ···················· | ···················· | ····················· | ···················· | ···················· | ···················· | | |
| I | 2015 hCoV-19/USA/WI-GMF-14557/2020\|EPI_ISL_547680\|2020- | ···················· | ···················· | ····················· | ···················· | ···················· | ···················· | | |
| I | 2016 hCoV-19/USA/WI-GMF-14585/2020\|EPI_ISL_547681\|2020- | ···················· | ·············A······ | ····················· | ···················· | ···················· | ···················· | | |
| I | 2017 hCoV-19/USA/WI-GMF-14586/2020\|EPI_ISL_547682\|2020- | ···················· | ···················· | ····················· | ···················· | ···················· | ···················· | | |
| I | 2018 hCoV-19/USA/WI-GMF-M00027/2020\|EPI_ISL_547683\|2020 | ···················· | ···················· | ····················· | ···················· | ···················· | ···················· | | |
| I | 2019 hCoV-19/USA/WI-GMF-14621/2020\|EPI_ISL_547685\|2020- | ···················· | ···················· | ····················· | ···················· | ···················· | ···················· | | |
| I | 2020 hCoV-19/USA/WI-GMF-14724/2020\|EPI_ISL_547687\|2020- | ···················· | ···················· | ····················· | ···················· | ···················· | ···················· | | |
| I | 2021 hCoV-19/USA/WI-GMF-14753/2020\|EPI_ISL_547688\|2020- | ···················· | ···················· | ····················· | ···················· | ···················· | ···················· | | |
| I | 2022 hCoV-19/USA/WI-GMF-14849/2020\|EPI_ISL_547689\|2020- | ···················· | ···················· | ····················· | ···················· | ···················· | ···················· | | |
| I | 2023 hCoV-19/USA/WI-GMF-14850/2020\|EPI_ISL_547690\|2020- | ···················· | ···················· | ····················· | ···················· | ···················· | ···················· | | |
| I | 2024 hCoV-19/USA/WI-GMF-14857/2020\|EPI_ISL_547691\|2020- | ···················· | ···················· | ····················· | ···················· | ···················· | ···················· | | |
| I | 2025 hCoV-19/USA/MN-GMF-14871/2020\|EPI_ISL_547692\|2020- | ···················· | ·············A······ | ····················· | ···················· | ···················· | ···················· | | |
| I | 2026 hCoV-19/USA/MN-GMF-14999/2020\|EPI_ISL_547693\|2020- | ···················· | ·············A······ | ····················· | ···················· | ···················· | ···················· | | |
| I | 2027 hCoV-19/USA/WI-GMF-15023/2020\|EPI_ISL_547694\|2020- | ···················· | ···················· | ····················· | ···················· | ···················· | ···················· | | |
| I | 2028 hCoV-19/USA/WI-GMF-15024/2020\|EPI_ISL_547695\|2020- | ···················· | ···················· | ····················· | ···················· | ···················· | ···················· | | |
| I | 2029 hCoV-19/USA/MN-GMF-15040/2020\|EPI_ISL_547696\|2020- | ···················· | ···················· | ····················· | ···················· | ···················· | ···················· | | |
| I | 2030 hCoV-19/USA/WI-GMF-15072/2020\|EPI_ISL_547697\|2020- | ···················· | ···················· | ····················· | ···················· | ···················· | ···················· | | |
| I | 2031 hCoV-19/USA/WI-GMF-15087/2020\|EPI_ISL_547698\|2020- | ···················· | ···················· | ····················· | ···················· | ···················· | ···················· | | |
| I | 2032 hCoV-19/Canada/ON-PHL-20-01296/2020\|EPI_ISL_548900 | ···················· | ···················· | ····················· | ···················· | ···················· | ···················· | | |
| I | 2033 hCoV-19/Canada/ON-PHL-20-01297/2020\|EPI_ISL_548901 | ···················· | ···················· | ····················· | ···················· | ···················· | ···················· | | |
| I | 2034 hCoV-19/Canada/ON-PHL-20-01298/2020\|EPI_ISL_548902 | ···················· | ···················· | ····················· | ···················· | ···················· | ···················· | | |
| I | 2035 hCoV-19/Canada/ON-PHL-20-01299/2020\|EPI_ISL_548903 | ···················· | ···················· | ····················· | ···················· | ···················· | ···················· | | |
| I | 2036 hCoV-19/Canada/ON-PHL-20-01300/2020\|EPI_ISL_548904 | ···················· | ···················· | ····················· | ···················· | NNN················· | ···················· | | 1 |
| I | 2037 hCoV-19/Canada/ON-PHL-20-01302/2020\|EPI_ISL_548905 | ···················· | ···················· | ····················· | ···················· | ···················· | ···················· | | |
| I | 2038 hCoV-19/Canada/ON-PHL-20-01303/2020\|EPI_ISL_548906 | ···················· | ···················· | ····················· | ···················· | ···················· | ···················· | | |
| I | 2039 hCoV-19/Canada/ON-PHL-20-01305/2020\|EPI_ISL_548908 | ···················· | ···················· | ····················· | ···················· | ···················· | ···················· | | |
| I | 2041 hCoV-19/Canada/ON-PHL-20-01307/2020\|EPI_ISL_548910 | ···················· | ···················· | ····················· | ···················· | ···················· | ···················· | | |
| I | 2042 hCoV-19/Canada/ON-PHL-20-01308/2020\|EPI_ISL_548911 | ···················· | ···················· | ····················· | ···················· | ···················· | ···················· | | |
| I | 2044 hCoV-19/Canada/ON-PHL-20-01312/2020\|EPI_ISL_548915 | ···················· | ···················· | ····················· | ···················· | ···················· | ···················· | | |
| I | 2045 hCoV-19/Canada/ON-PHL-20-01317/2020\|EPI_ISL_548920 | ···················· | ···················· | ····················· | ···················· | ···················· | ···················· | | |
| I | 2046 hCoV-19/Canada/ON-PHL-20-01318/2020\|EPI_ISL_548921 | ···················· | ···················· | ····················· | ···················· | ···················· | ···················· | | |
| I | 2047 hCoV-19/Canada/ON-PHL-20-01319/2020\|EPI_ISL_548922 | ···················· | ···················· | ····················· | ···················· | ···················· | ···················· | | |
| I | 2048 hCoV-19/Canada/ON-PHL-20-01320/2020\|EPI_ISL_548923 | ···················· | ···················· | ····················· | ···················· | ···················· | ···················· | | |
| I | 2049 hCoV-19/USA/TX-HMH-5148/2020\|EPI_ISL_544558\|2020-0 | ···················· | ···················· | ····················· | ···················· | ···················· | ···················· | | |
| I | 2050 hCoV-19/USA/TX-HMH-5161/2020\|EPI_ISL_544568\|2020-0 | ···················· | ···················· | ····················· | ···················· | ···················· | ···················· | | |
| I | 2051 hCoV-19/USA/TX-HMH-5162/2020\|EPI_ISL_544569\|2020-0 | ···················· | ···················· | ····················· | ···················· | ···················· | ···················· | | |
| I | 2054 hCoV-19/USA/TX-HMH-5166/2020\|EPI_ISL_544572\|2020-0 | ···················· | ···················· | ····················· | ···················· | ···················· | ···················· | | |
| I | 2055 hCoV-19/USA/TX-HMH-5167/2020\|EPI_ISL_544573\|2020-0 | ···················· | ···················· | ····················· | ···················· | ···················· | ··N················· | | |
| I | 2056 hCoV-19/USA/TX-HMH-5168/2020\|EPI_ISL_544574\|2020-0 | ···················· | ···················· | ····················· | ···················· | ···················· | ···················· | | |
| I | 2057 hCoV-19/USA/TX-HMH-5174/2020\|EPI_ISL_544575\|2020-0 | ···················· | ···················· | ····················· | ···················· | ···················· | ···················· | | |
| I | 2058 hCoV-19/USA/TX-HMH-5176/2020\|EPI_ISL_544576\|2020-0 | ···················· | ···················· | ····················· | ···················· | ···················· | ···················· | | |
| I | 2059 hCoV-19/USA/TX-HMH-5179/2020\|EPI_ISL_544578\|2020-0 | ···················· | ···················· | ····················· | ···················· | ···················· | ···················· | | |
| I | 2060 hCoV-19/USA/TX-HMH-518/2020\|EPI_ISL_544579\|2020-04 | ···················· | ···················· | ····················· | ···················· | ···················· | ···················· | | |
| I | 2061 hCoV-19/USA/TX-HMH-5181/2020\|EPI_ISL_544580\|2020-0 | ···················· | ···················· | ····················· | ···················· | ···················· | ···················· | | |
| I | 2065 hCoV-19/USA/TX-HMH-519/2020\|EPI_ISL_544585\|2020-04 | ···················· | ···················· | ····················· | ···················· | ···················· | ···················· | | |
| I | 2067 hCoV-19/USA/TX-HMH-5192/2020\|EPI_ISL_544587\|2020-0 | ···················· | ···················· | ····················· | ···················· | ··············NN·N·· | ·······N············ | 1 | 1 |
| I | 2068 hCoV-19/USA/TX-HMH-520/2020\|EPI_ISL_544588\|2020-04 | ···················· | ···················· | ····················· | ···················· | ···················· | ···················· | | |
| I | 2071 hCoV-19/USA/TX-HMH-521/2020\|EPI_ISL_544595\|2020-04 | ···················· | ···················· | ····················· | ···················· | ···················· | ···················· | | |
| I | 2072 hCoV-19/USA/TX-HMH-5210/2020\|EPI_ISL_544596\|2020-0 | ···················· | ···················· | ····················· | ···················· | ···················· | ···················· | | |
| I | 2075 hCoV-19/USA/TX-HMH-5217/2020\|EPI_ISL_544601\|2020-0 | ···················· | ···················· | ····················· | ···················· | ···················· | ···················· | | |
| I | 2076 hCoV-19/USA/TX-HMH-5218/2020\|EPI_ISL_544602\|2020-0 | ···················· | ···················· | ····················· | ···················· | ···················· | NN·················· | | |

| | | | | | | | | | |
|---|---|---|---|---|---|---|---|---|---|
| I | 2078 | hCoV-19/USA/TX-HMH-522/2020\|EPI_ISL_544604\|2020-04 | ···················· | ···················· | ···················· | ···················· | ···················· | ···················· | |
| I | 2080 | hCoV-19/USA/TX-HMH-5222/2020\|EPI_ISL_544607\|2020-0 | ···················· | ···················· | ···················· | ···················· | ···················· | ···················· | |
| I | 2082 | hCoV-19/USA/TX-HMH-5232/2020\|EPI_ISL_544611\|2020-0 | ···················· | ···················· | ···················· | ···················· | ···················· | ···················· | |
| I | 2084 | hCoV-19/USA/TX-HMH-5235/2020\|EPI_ISL_544613\|2020-0 | ···················· | ···················· | ···················· | ···················· | ···················· | ···················· | |
| I | 2089 | hCoV-19/USA/TX-HMH-5243/2020\|EPI_ISL_544620\|2020-0 | ···················· | ···················· | ···················· | ···················· | ···················· | ···················· | |
| I | 2091 | hCoV-19/USA/TX-HMH-5246/2020\|EPI_ISL_544623\|2020-0 | ···················· | ·············A······ | ···················· | ···················· | ···················· | ···················· | |
| I | 2095 | hCoV-19/USA/TX-HMH-5256/2020\|EPI_ISL_544630\|2020-0 | ···················· | ···················· | ···················· | ···················· | ···················· | ···················· | |
| I | 2098 | hCoV-19/USA/TX-HMH-5263/2020\|EPI_ISL_544635\|2020-0 | ···················· | ···················· | ···················· | ···················· | ···················· | ···················· | |
| I | 2099 | hCoV-19/USA/TX-HMH-5268/2020\|EPI_ISL_544639\|2020-0 | ···················· | ···················· | ···················· | ···················· | ···················· | ···················· | |
| I | 2101 | hCoV-19/USA/TX-HMH-5271/2020\|EPI_ISL_544641\|2020-0 | ···················· | ···················· | ···················· | ···················· | ···················· | ···················· | |
| I | 2102 | hCoV-19/USA/TX-HMH-5272/2020\|EPI_ISL_544642\|2020-0 | ···················· | ···················· | ···················· | ···················· | ···················· | ···················· | |
| I | 2104 | hCoV-19/USA/TX-HMH-2001/2020\|EPI_ISL_545060\|2020-0 | ···················· | ······A············· | ···················· | ···················· | ···················· | ···················· | |
| I | 2105 | hCoV-19/USA/TX-HMH-2002/2020\|EPI_ISL_545061\|2020-0 | ···················· | ···················· | ···················· | ···················· | ···················· | ···················· | |
| I | 2106 | hCoV-19/USA/TX-HMH-2003/2020\|EPI_ISL_545062\|2020-0 | ···················· | ······A············· | ···················· | ···················· | ···················· | ···················· | |
| I | 2107 | hCoV-19/USA/TX-HMH-2004/2020\|EPI_ISL_545063\|2020-0 | ···················· | ···················· | ···················· | ···················· | ···················· | ···················· | |
| I | 2108 | hCoV-19/USA/TX-HMH-2005/2020\|EPI_ISL_545064\|2020-0 | ···················· | ···················· | ···················· | ···················· | ···················· | ···················· | |
| I | 2109 | hCoV-19/USA/TX-HMH-2008/2020\|EPI_ISL_545065\|2020-0 | ···················· | ······A············· | ···················· | ···················· | ···················· | ···················· | |
| I | 2110 | hCoV-19/USA/TX-HMH-2009/2020\|EPI_ISL_545066\|2020-0 | ···················· | ··············C····· | ···················· | ···················· | ···················· | ···················· | |
| I | 2111 | hCoV-19/USA/TX-HMH-2013/2020\|EPI_ISL_545068\|2020-0 | ···················· | ···················· | ···················· | ···················· | ···················· | ···················· | |
| I | 2112 | hCoV-19/USA/TX-HMH-2015/2020\|EPI_ISL_545069\|2020-0 | ···················· | ···················· | ···················· | ···················· | ···················· | ···················· | |
| I | 2113 | hCoV-19/USA/TX-HMH-2019/2020\|EPI_ISL_545070\|2020-0 | ···················· | ···················· | ···················· | ···················· | ···················· | ···················· | |
| I | 2114 | hCoV-19/USA/TX-HMH-2021/2020\|EPI_ISL_545071\|2020-0 | ···················· | ···················· | ···················· | ···················· | ···················· | ···················· | |
| I | 2115 | hCoV-19/USA/TX-HMH-2022/2020\|EPI_ISL_545072\|2020-0 | ···················· | ···················· | ···················· | ···················· | ···················· | ···················· | |
| I | 2116 | hCoV-19/USA/TX-HMH-2023/2020\|EPI_ISL_545073\|2020-0 | ···················· | ···················· | ···················· | ···················· | ···················· | ···················· | |
| I | 2117 | hCoV-19/USA/TX-HMH-2030/2020\|EPI_ISL_545075\|2020-0 | ···················· | ···················· | ···················· | ···················· | ···················· | ···················· | |
| I | 2118 | hCoV-19/USA/TX-HMH-2034/2020\|EPI_ISL_545076\|2020-0 | ···················· | ··············C····· | ···················· | ···················· | ···················· | ···················· | |
| I | 2119 | hCoV-19/USA/TX-HMH-2038/2020\|EPI_ISL_545078\|2020-0 | ···················· | ···················· | ···················· | ···················· | ···················· | ···················· | |
| I | 2120 | hCoV-19/USA/TX-HMH-2040/2020\|EPI_ISL_545079\|2020-0 | ···················· | ···················· | ···················· | ···················· | ···················· | ···················· | |
| I | 2121 | hCoV-19/USA/TX-HMH-2041/2020\|EPI_ISL_545080\|2020-0 | ···················· | ···················· | ···················· | ···················· | ···················· | ···················· | |
| I | 2122 | hCoV-19/USA/TX-HMH-2042/2020\|EPI_ISL_545081\|2020-0 | ···················· | ···················· | ···················· | ···················· | ···················· | ···················· | |
| I | 2123 | hCoV-19/USA/TX-HMH-2043/2020\|EPI_ISL_545082\|2020-0 | ···················· | ···················· | ···················· | ···················· | ···················· | ···················· | |
| I | 2124 | hCoV-19/USA/TX-HMH-2044/2020\|EPI_ISL_545083\|2020-0 | ···················· | ···················· | ···················· | ···················· | ···················· | ···················· | |
| I | 2125 | hCoV-19/USA/TX-HMH-2045/2020\|EPI_ISL_545084\|2020-0 | ···················· | ···················· | ···················· | ···················· | ···················· | ···················· | |
| I | 2126 | hCoV-19/USA/TX-HMH-2046/2020\|EPI_ISL_545085\|2020-0 | ···················· | ···················· | ···················· | ···················· | ···················· | ···················· | |
| I | 2127 | hCoV-19/USA/TX-HMH-2050/2020\|EPI_ISL_545087\|2020-0 | ···················· | ···················· | ···················· | ···················· | ···················· | ···················· | |
| I | 2128 | hCoV-19/USA/TX-HMH-2051/2020\|EPI_ISL_545088\|2020-0 | ···················· | ···················· | ···················· | ···················· | ···················· | ···················· | |
| I | 2129 | hCoV-19/USA/TX-HMH-2057/2020\|EPI_ISL_545093\|2020-0 | ···················· | ···················· | ···················· | ···················· | ···················· | ···················· | |
| I | 2130 | hCoV-19/USA/TX-HMH-2058/2020\|EPI_ISL_545094\|2020-0 | ···················· | ···················· | ···················· | ···················· | ········N·····N····· | NN·A················ | 1 |
| I | 2131 | hCoV-19/USA/TX-HMH-2061/2020\|EPI_ISL_545095\|2020-0 | ···················· | ···················· | ···················· | ···················· | ···················· | ···················· | |
| I | 2132 | hCoV-19/USA/TX-HMH-2062/2020\|EPI_ISL_545096\|2020-0 | ···················· | ···················· | ···················· | ···················· | ···················· | ···················· | |
| I | 2133 | hCoV-19/USA/TX-HMH-2063/2020\|EPI_ISL_545097\|2020-0 | ···················· | ···················· | ···················· | ···················· | ···················· | ···················· | |
| I | 2134 | hCoV-19/USA/TX-HMH-872/2020\|EPI_ISL_545889\|2020-04 | ···················· | ···················· | ···················· | ···················· | ···················· | ···················· | |
| I | 2135 | hCoV-19/USA/TX-HMH-886/2020\|EPI_ISL_545899\|2020-04 | ···················· | ···················· | ···················· | ···················· | ···················· | ···················· | |
| I | 2136 | hCoV-19/USA/TX-HMH-887/2020\|EPI_ISL_545900\|2020-04 | ···················· | ···················· | ···················· | ···················· | ···················· | ···················· | |
| I | 2137 | hCoV-19/USA/TX-HMH-888/2020\|EPI_ISL_545901\|2020-04 | ···················· | ···················· | ···················· | ···················· | ···················· | ···················· | |
| I | 2138 | hCoV-19/USA/TX-HMH-889/2020\|EPI_ISL_545902\|2020-04 | ···················· | ···················· | ···················· | ···················· | ···················· | ···················· | |
| I | 2139 | hCoV-19/USA/TX-HMH-890/2020\|EPI_ISL_545903\|2020-04 | ···················· | ···················· | ···················· | ···················· | ···················· | ···················· | |
| I | 2140 | hCoV-19/USA/TX-HMH-893/2020\|EPI_ISL_545905\|2020-04 | ···················· | ···················· | ···················· | ···················· | ···················· | ···················· | |
| I | 2141 | hCoV-19/USA/TX-HMH-894/2020\|EPI_ISL_545906\|2020-04 | ···················· | ···················· | ···················· | ···················· | ···················· | ···················· | |
| I | 2142 | hCoV-19/USA/TX-HMH-895/2020\|EPI_ISL_545907\|2020-04 | ···················· | ···················· | ···················· | ···················· | ···················· | ···················· | |
| I | 2143 | hCoV-19/USA/TX-HMH-896/2020\|EPI_ISL_545908\|2020-04 | ···················· | ···················· | ···················· | ···················· | ···················· | ···················· | |
| I | 2144 | hCoV-19/USA/TX-HMH-906/2020\|EPI_ISL_545909\|2020-04 | ···················· | ···················· | ···················· | ···················· | ···················· | ···················· | |
| I | 2145 | hCoV-19/USA/TX-HMH-909/2020\|EPI_ISL_545911\|2020-04 | ···················· | ···················· | ···················· | ···················· | ···················· | ···················· | |
| I | 2146 | hCoV-19/USA/TX-HMH-911/2020\|EPI_ISL_545912\|2020-04 | ···················· | ···················· | ···················· | ···················· | ···················· | ···················· | |
| I | 2147 | hCoV-19/USA/TX-HMH-912/2020\|EPI_ISL_545913\|2020-04 | ···················· | ···················· | ···················· | ···················· | ···················· | ···················· | |
| I | 2149 | hCoV-19/USA/TX-HMH-914/2020\|EPI_ISL_545915\|2020-04 | ···················· | ···················· | ···················· | ···················· | ···················· | ···················· | |
| I | 2150 | hCoV-19/USA/TX-HMH-915/2020\|EPI_ISL_545916\|2020-04 | ···················· | ···················· | ···················· | ···················· | ···················· | ···················· | |
| I | 2152 | hCoV-19/USA/TX-HMH-920/2020\|EPI_ISL_545919\|2020-04 | ···················· | ···················· | ···················· | ···················· | ···················· | ···················· | |
| I | 2153 | hCoV-19/USA/TX-HMH-921/2020\|EPI_ISL_545920\|2020-04 | ···················· | ···················· | ···················· | ···················· | ···················· | ···················· | |
| I | 2154 | hCoV-19/USA/TX-HMH-922/2020\|EPI_ISL_545921\|2020-04 | ···················· | ···················· | ···················· | ···················· | ···················· | ···················· | |
| I | 2155 | hCoV-19/USA/TX-HMH-924/2020\|EPI_ISL_545922\|2020-04 | ···················· | ···················· | ···················· | ···················· | ···················· | ···················· | |
| I | 2157 | hCoV-19/USA/TX-HMH-929/2020\|EPI_ISL_545924\|2020-04 | ···················· | ···················· | ···················· | ···················· | ···················· | ···················· | |
| I | 2158 | hCoV-19/USA/TX-HMH-934/2020\|EPI_ISL_545926\|2020-04 | ···················· | ···················· | ···················· | ···················· | ···················· | ···················· | |
| I | 2159 | hCoV-19/USA/TX-HMH-944/2020\|EPI_ISL_545927\|2020-04 | ···················· | ···················· | ···················· | ···················· | ···················· | ···················· | |
| I | 2160 | hCoV-19/USA/TX-HMH-948/2020\|EPI_ISL_545928\|2020-04 | ···················· | ···················· | ···················· | ···················· | ···················· | ···················· | |
| I | 2161 | hCoV-19/USA/TX-HMH-955/2020\|EPI_ISL_545929\|2020-04 | ···················· | ···················· | ···················· | ···················· | ···················· | ···················· | |
| I | 2163 | hCoV-19/USA/TX-HMH-957/2020\|EPI_ISL_545931\|2020-04 | ···················· | ···················· | ···················· | ···················· | ···················· | ···················· | |
| I | 2164 | hCoV-19/USA/TX-HMH-958/2020\|EPI_ISL_545932\|2020-04 | ···················· | ···················· | ···················· | ···················· | ···················· | ···················· | |
| I | 2165 | hCoV-19/USA/TX-HMH-959/2020\|EPI_ISL_545933\|2020-04 | ···················· | ···················· | ···················· | ···················· | ···················· | ···················· | |
| I | 2166 | hCoV-19/USA/TX-HMH-960/2020\|EPI_ISL_545934\|2020-04 | ···················· | ···················· | ···················· | ···················· | ···················· | ···················· | |
| I | 2167 | hCoV-19/USA/TX-HMH-961/2020\|EPI_ISL_545935\|2020-04 | ···················· | ···················· | ···················· | ···················· | ···················· | ···················· | |
| I | 2168 | hCoV-19/USA/TX-HMH-962/2020\|EPI_ISL_545936\|2020-04 | ···················· | ···················· | ···················· | ···················· | ···················· | ···················· | |
| I | 2169 | hCoV-19/USA/TX-HMH-967/2020\|EPI_ISL_545937\|2020-04 | ···················· | ···················· | ···················· | ···················· | ···················· | ···················· | |
| I | 2170 | hCoV-19/USA/TX-HMH-969/2020\|EPI_ISL_545938\|2020-04 | ···················· | ···················· | ···················· | ···················· | ···················· | ···················· | |
| I | 2171 | hCoV-19/USA/TX-HMH-970/2020\|EPI_ISL_545939\|2020-04 | ···················· | ···················· | ···················· | ···················· | ···················· | ···················· | |
| I | 2172 | hCoV-19/USA/TX-HMH-973/2020\|EPI_ISL_545940\|2020-04 | ···················· | ···················· | ···················· | ···················· | ···················· | ···················· | |
| I | 2173 | hCoV-19/USA/TX-HMH-974/2020\|EPI_ISL_545941\|2020-04 | ···················· | ···················· | ···················· | ···················· | ···················· | ···················· | |
| I | 2174 | hCoV-19/USA/TX-HMH-975/2020\|EPI_ISL_545942\|2020-04 | ···················· | ···················· | ···················· | ···················· | ···················· | ···················· | |
| I | 2176 | hCoV-19/USA/TX-HMH-983/2020\|EPI_ISL_545945\|2020-04 | ···················· | ···················· | ···················· | ···················· | ···················· | ···················· | |
| I | 2177 | hCoV-19/USA/TX-HMH-984/2020\|EPI_ISL_545946\|2020-04 | ···················· | ···················· | ···················· | ···················· | ···················· | ···················· | |

| | | | | | | | |
|---|---|---|---|---|---|---|---|
| I | 2178 hCoV-19/USA/TX-HMH-985/2020\|EPI_ISL_545947\|2020-04 | ···················· | ···················· | ···················· | ···················· | ···················· | ···················· |
| I | 2179 hCoV-19/USA/TX-HMH-986/2020\|EPI_ISL_545948\|2020-04 | ···················· | ···················· | ···················· | ···················· | ···················· | ···················· |
| I | 2180 hCoV-19/USA/TX-HMH-992/2020\|EPI_ISL_545949\|2020-04 | ···················· | ···················· | ···················· | ···················· | ···················· | ···················· |
| I | 2181 hCoV-19/USA/TX-HMH-994/2020\|EPI_ISL_545951\|2020-04 | ···················· | ···················· | ···················· | ···················· | ···················· | ···················· |
| I | 2182 hCoV-19/USA/TX-HMH-995/2020\|EPI_ISL_545952\|2020-04 | ···················· | ···················· | ···················· | ···················· | ···················· | ···················· |
| I | 2183 hCoV-19/USA/TX-HMH-996/2020\|EPI_ISL_545953\|2020-04 | ···················· | ···················· | ···················· | ···················· | ···················· | ···················· |
| I | 2184 hCoV-19/Czech Republic/NRL_7918/2020\|EPI_ISL_54595 | ···················· | ···················· | ···················· | ···················· | ···················· | ···················· |
| I | 2185 hCoV-19/Czech Republic/NRL_7942/2020\|EPI_ISL_54595 | ···················· | ···················· | ···················· | ···················· | ···················· | ···················· |
| I | 2186 hCoV-19/Czech Republic/NRL-7944/2020\|EPI_ISL_54595 | ···················· | ···················· | ···················· | ···················· | ···················· | ···················· |
| I | 2188 hCoV-19/USA/TX-HMH-6387/2020\|EPI_ISL_545960\|2020-0 | ···················· | ·············A······ | ···················· | ···················· | ···················· | ···················· |
| I | 2189 hCoV-19/USA/TX-HMH-6388/2020\|EPI_ISL_545961\|2020-0 | ···················· | ···················· | ···················· | ···················· | ···················· | ···················· |
| I | 2190 hCoV-19/USA/TX-HMH-6390/2020\|EPI_ISL_545963\|2020-0 | ···················· | ···················· | ···················· | ···················· | ···················· | ···················· |
| I | 2191 hCoV-19/USA/TX-HMH-6391/2020\|EPI_ISL_545964\|2020-0 | ···················· | ···················· | ···················· | ···················· | ···················· | ···················· |
| I | 2192 hCoV-19/USA/TX-HMH-6392/2020\|EPI_ISL_545965\|2020-0 | ···················· | ···················· | ···················· | ···················· | ···················· | ···················· |
| I | 2193 hCoV-19/USA/TX-HMH-6394/2020\|EPI_ISL_545967\|2020-0 | ···················· | ···················· | ···················· | ···················· | ···················· | ···················· |
| I | 2194 hCoV-19/USA/TX-HMH-6395/2020\|EPI_ISL_545968\|2020-0 | ···················· | ···················· | ···················· | ···················· | ···················· | ···················· |
| I | 2196 hCoV-19/USA/TX-HMH-6401/2020\|EPI_ISL_545973\|2020-0 | ···················· | ·············A······ | ···················· | ···················· | ···················· | ···N················ |
| I | 2198 hCoV-19/USA/TX-HMH-6865/2020\|EPI_ISL_546300\|2020-0 | ···················· | ···················· | ···················· | ···················· | ···················· | ···················· |
| I | 2200 hCoV-19/USA/TX-HMH-6867/2020\|EPI_ISL_546302\|2020-0 | ···················· | ···················· | ···················· | ···················· | ···················· | ···················· |
| I | 2202 hCoV-19/USA/TX-HMH-687/2020\|EPI_ISL_546305\|2020-04 | ···················· | ···················· | ···················· | ···················· | ···················· | ···················· |
| I | 2203 hCoV-19/USA/TX-HMH-6963/2020\|EPI_ISL_546390\|2020-0 | ···················· | ···················· | ···················· | ···················· | ···················· | ···················· |
| I | 2205 hCoV-19/USA/TX-HMH-6965/2020\|EPI_ISL_546392\|2020-0 | ···················· | ···················· | ···················· | ···················· | ···················· | ···················· |
| I | 2207 hCoV-19/USA/TX-HMH-6971/2020\|EPI_ISL_546397\|2020-0 | ···················· | ·············A······ | ···················· | ···················· | ···················· | ···················· |
| I | 2209 hCoV-19/Australia/NSW935/2020\|EPI_ISL_547600\|2020- | ···················· | ···················· | ···················· | ···················· | ···················· | ···················· |
| I | 2210 hCoV-19/Australia/NSW936/2020\|EPI_ISL_547601\|2020- | ···················· | ···················· | ···················· | ···················· | ···················· | ···················· |
| I | 2211 hCoV-19/USA/WI-GMF-09851/2020\|EPI_ISL_547602\|2020- | ···················· | ···················· | ···················· | ···················· | ···················· | ···················· |
| I | 2212 hCoV-19/USA/WI-GMF-09884/2020\|EPI_ISL_547603\|2020- | ···················· | ·A·················· | ···················· | ···················· | ···················· | ···················· |
| I | 2213 hCoV-19/USA/IA-GMF-09911/2020\|EPI_ISL_547604\|2020- | ···················· | ···················· | ···················· | ···················· | ···················· | ···················· |
| I | 2214 hCoV-19/USA/WI-GMF-09931/2020\|EPI_ISL_547605\|2020- | ···················· | ···················· | ···················· | ···················· | ···················· | ···················· |
| I | 2215 hCoV-19/USA/WI-GMF-09934/2020\|EPI_ISL_547606\|2020- | ···················· | ···················· | ···················· | ···················· | ···················· | ···················· |
| I | 2216 hCoV-19/USA/IA-GMF-10147/2020\|EPI_ISL_547607\|2020- | ···················· | ···················· | ···················· | ···················· | ···················· | ···················· |
| I | 2217 hCoV-19/USA/WI-GMF-10366/2020\|EPI_ISL_547608\|2020- | ···················· | ···················· | ···················· | ···················· | ···················· | ···················· |
| I | 2218 hCoV-19/USA/WI-GMF-10451/2020\|EPI_ISL_547609\|2020- | ···················· | ···················· | ···················· | ···················· | ···················· | ···················· |
| I | 2219 hCoV-19/USA/WI-GMF-10571/2020\|EPI_ISL_547610\|2020- | ···················· | ······A············· | ···················· | ···················· | ···················· | ···················· |
| I | 2220 hCoV-19/USA/WI-GMF-10757/2020\|EPI_ISL_547611\|2020- | ···················· | ···················· | ···················· | ···················· | ···················· | ···················· |
| I | 2221 hCoV-19/USA/WI-GMF-10762/2020\|EPI_ISL_547612\|2020- | ···················· | ···················· | ···················· | ···················· | ···················· | ···················· |
| I | 2222 hCoV-19/USA/WI-GMF-10809/2020\|EPI_ISL_547613\|2020- | ···················· | ·A·················· | ···················· | ···················· | K··················· | ···················· |
| I | 2223 hCoV-19/USA/WI-GMF-10819/2020\|EPI_ISL_547614\|2020- | ···················· | ·A·················· | ···················· | ···················· | ···················· | ···················· |
| I | 2224 hCoV-19/USA/WI-GMF-10838/2020\|EPI_ISL_547615\|2020- | ···················· | ···················· | ···················· | ···················· | ···················· | ···················· |
| I | 2225 hCoV-19/USA/WI-GMF-10854/2020\|EPI_ISL_547616\|2020- | ···················· | ···················· | ···················· | ···················· | ···················· | ···················· |
| I | 2226 hCoV-19/USA/WI-GMF-10885/2020\|EPI_ISL_547617\|2020- | ···················· | ···················· | ···················· | ···················· | ···················· | ···················· |
| I | 2227 hCoV-19/USA/WI-GMF-10889/2020\|EPI_ISL_547618\|2020- | ···················· | ···················· | ···················· | ···················· | ···················· | ···················· |
| I | 2228 hCoV-19/USA/WI-GMF-10900/2020\|EPI_ISL_547619\|2020- | ···················· | ···················· | ···················· | ···················· | ···················· | ···················· |
| I | 2229 hCoV-19/USA/WI-GMF-10913/2020\|EPI_ISL_547620\|2020- | ···················· | ···················· | ···················· | ···················· | ···················· | ···················· |
| I | 2230 hCoV-19/USA/WI-GMF-10983/2020\|EPI_ISL_547621\|2020- | ···················· | ···················· | ···················· | ···················· | ···················· | ···················· |
| I | 2231 hCoV-19/USA/WI-GMF-11002/2020\|EPI_ISL_547622\|2020- | ···················· | ···················· | ···················· | ···················· | ···················· | ···················· |
| I | 2232 hCoV-19/USA/WI-GMF-11060/2020\|EPI_ISL_547623\|2020- | ···················· | ·············A······ | ···················· | ···················· | ···················· | ···················· |
| I | 2233 hCoV-19/USA/WI-GMF-11066/2020\|EPI_ISL_547624\|2020- | ···················· | ···················· | ···················· | ···················· | ···················· | ···················· |
| I | 2234 hCoV-19/USA/WI-GMF-11146/2020\|EPI_ISL_547626\|2020- | ···················· | ···················· | ···················· | ···················· | ···················· | ···················· |
| I | 2235 hCoV-19/USA/IA-GMF-11334/2020\|EPI_ISL_547627\|2020- | ···················· | ···················· | ···················· | ···················· | ···················· | ···················· |
| I | 2236 hCoV-19/USA/WI-GMF-11415/2020\|EPI_ISL_547628\|2020- | ···················· | ···················· | ···················· | ···················· | ···················· | ···················· |
| I | 2237 hCoV-19/USA/WI-GMF-11426/2020\|EPI_ISL_547629\|2020- | ···················· | ···················· | ···················· | ···················· | ···················· | ···················· |
| I | 2238 hCoV-19/USA/WI-GMF-11475/2020\|EPI_ISL_547630\|2020- | ···················· | ···················· | ···················· | ···················· | ···················· | ···················· |
| I | 2239 hCoV-19/USA/WI-GMF-11582/2020\|EPI_ISL_547631\|2020- | ···················· | ···················· | ···················· | ···················· | ···················· | ···················· |
| I | 2240 hCoV-19/USA/WI-GMF-11750/2020\|EPI_ISL_547632\|2020- | ···················· | ···················· | ···················· | ···················· | ···················· | ···················· |
| I | 2241 hCoV-19/USA/WI-GMF-11818/2020\|EPI_ISL_547633\|2020- | ···················· | ···················· | ···················· | ···················· | ···················· | ···················· |
| I | 2242 hCoV-19/USA/WI-GMF-11874/2020\|EPI_ISL_547634\|2020- | ···················· | ···················· | ···················· | ···················· | ···················· | ···················· |
| I | 2243 hCoV-19/USA/WI-GMF-11890/2020\|EPI_ISL_547635\|2020- | ···T················ | ·A·················· | ···················· | ···················· | ···················· | ···················· |
| I | 2244 hCoV-19/USA/WI-GMF-11927/2020\|EPI_ISL_547636\|2020- | ···················· | ···················· | ···················· | ···················· | ···················· | ···················· |
| I | 2245 hCoV-19/New Zealand/20VR3916/2020\|EPI_ISL_548100\|2 | ···················· | ···················· | ···················· | ···················· | ···················· | ···················· |
| I | 2246 hCoV-19/New Zealand/20VR3919/2020\|EPI_ISL_548101\|2 | ···················· | ···················· | ···················· | ···················· | ···················· | ···················· |
| I | 2247 hCoV-19/New Zealand/20CV0063/2020\|EPI_ISL_548104\|2 | ···················· | ·A·················· | ···················· | ···················· | ···················· | ···A················ |
| I | 2248 hCoV-19/New Zealand/20CV0071/2020\|EPI_ISL_548106\|2 | ···················· | ···················· | ···················· | ···················· | ···················· | ···················· |
| I | 2250 hCoV-19/New Zealand/20CV0202/2020\|EPI_ISL_548110\|2 | ···················· | ·A·················· | ···················· | ···················· | ···················· | ···A················ |
| I | 2251 hCoV-19/New Zealand/20CV0203/2020\|EPI_ISL_548111\|2 | ···················· | ·A·················· | ···················· | ···················· | ···················· | ···A················ |
| I | 2252 hCoV-19/New Zealand/20VR2095/2020\|EPI_ISL_548123\|2 | ···················· | ···················· | ···················· | ···················· | ···················· | ···················· |
| I | 2253 hCoV-19/Switzerland/GE-SNRCI-3674/2020\|EPI_ISL_548 | ···················· | ···················· | ···················· | ···················· | ···················· | ···················· |
| I | 2254 hCoV-19/Switzerland/GE-SNRCI-3780/2020\|EPI_ISL_548 | ···················· | ···················· | ···················· | ···················· | ···················· | ···················· |
| I | 2255 hCoV-19/Switzerland/GE-SNRCI-5671/2020\|EPI_ISL_548 | ···················· | ···················· | ···················· | ···················· | ···················· | ···················· |
| I | 2256 hCoV-19/Switzerland/GE-SNRCI-6128/2020\|EPI_ISL_548 | ···················· | ···················· | ···················· | ···················· | ···················· | ···················· |
| I | 2257 hCoV-19/Switzerland/GE-SNRCI-6350/2020\|EPI_ISL_548 | ···················· | ···················· | ···················· | ···················· | ···················· | ···················· |
| I | 2258 hCoV-19/Switzerland/GE-SNRCI-6448/2020\|EPI_ISL_548 | ···················· | ···················· | ···················· | ···················· | ···················· | ···················· |
| I | 2259 hCoV-19/Switzerland/GE-SNRCI-6477/2020\|EPI_ISL_548 | ···················· | ···················· | ···················· | ···················· | ···················· | ···················· |
| I | 2260 hCoV-19/Switzerland/GE-SNRCI-6699/2020\|EPI_ISL_548 | ···················· | ···················· | ···················· | ···················· | ···················· | ···················· |
| I | 2261 hCoV-19/Switzerland/GE-SNRCI-6763/2020\|EPI_ISL_548 | ···················· | ···················· | ···················· | ···················· | ···················· | ···················· |
| I | 2262 hCoV-19/Switzerland/GE-SNRCI-6852/2020\|EPI_ISL_548 | ···················· | ·A·················· | ···················· | ···················· | ···················· | ···················· |
| I | 2263 hCoV-19/Switzerland/GE-SNRCI-6928/2020\|EPI_ISL_548 | ···················· | ···················· | ···················· | ···················· | ···················· | ···················· |
| I | 2264 hCoV-19/Switzerland/GE-SNRCI-6955/2020\|EPI_ISL_548 | ···················· | ···················· | ···················· | ···················· | ···················· | ···················· |
| I | 2265 hCoV-19/Switzerland/GE-SNRCI-7014/2020\|EPI_ISL_548 | ···················· | ···················· | ···················· | ···················· | ···················· | ···················· |
| I | 2266 hCoV-19/Switzerland/GE-SNRCI-7058/2020\|EPI_ISL_548 | ···················· | ···················· | ···················· | ···················· | ···················· | ···················· |

| | | | | | | | | |
|---|---|---|---|---|---|---|---|---|
| I | 2267 hCoV-19/Switzerland/GE-SNRCI-7072/2020\|EPI_ISL_548 | ···················· | ···················· | ···················· | ···················· | ···················· | ···················· | |
| I | 2268 hCoV-19/Switzerland/GE-SNRCI-7201/2020\|EPI_ISL_548 | ···················· | ···················· | ···················· | ···················· | ···················· | ···················· | |
| I | 2269 hCoV-19/Switzerland/GE-SNRCI-7230/2020\|EPI_ISL_548 | ···················· | ···················· | ···················· | ···················· | ···················· | ···················· | |
| I | 2270 hCoV-19/Switzerland/GE-SNRCI-7310/2020\|EPI_ISL_548 | ···················· | ·A·················· | ···················· | ···················· | ···················· | ···················· | |
| I | 2271 hCoV-19/Switzerland/GE-SNRCI-7313/2020\|EPI_ISL_548 | ···················· | ···················· | ···················· | ···················· | ···················· | ···················· | |
| I | 2272 hCoV-19/Switzerland/GE-SNRCI-7347/2020\|EPI_ISL_548 | ···················· | ···················· | ···················· | ···················· | ···················· | ···················· | |
| I | 2273 hCoV-19/Switzerland/GE-SNRCI-7405/2020\|EPI_ISL_548 | ···················· | ···················· | ···················· | ···················· | ···················· | ···················· | |
| I | 2274 hCoV-19/Switzerland/GE-SNRCI-7419/2020\|EPI_ISL_548 | ···················· | ···················· | ···················· | ···················· | ···················· | ···················· | |
| I | 2275 hCoV-19/Switzerland/GE-SNRCI-7474/2020\|EPI_ISL_548 | ···················· | ···················· | ···················· | ···················· | ···················· | ···················· | |
| I | 2276 hCoV-19/Switzerland/GE-SNRCI-7517/2020\|EPI_ISL_548 | ···················· | ···················· | ···················· | ···················· | ···················· | ···················· | |
| I | 2277 hCoV-19/Switzerland/GE-SNRCI-7536/2020\|EPI_ISL_548 | ···················· | ···················· | ···················· | ···················· | ···················· | ···················· | |
| I | 2278 hCoV-19/Switzerland/GE-SNRCI-7548/2020\|EPI_ISL_548 | ···················· | ···················· | ···················· | ···················· | ···················· | ···················· | |
| I | 2279 hCoV-19/Switzerland/GE-SNRCI-7582/2020\|EPI_ISL_548 | ···················· | ···················· | ···················· | ···················· | ···················· | ···················· | |
| I | 2280 hCoV-19/Switzerland/GE-SNRCI-7699/2020\|EPI_ISL_548 | ···················· | ···················· | ···················· | ···················· | ···················· | ···················· | |
| I | 2281 hCoV-19/Switzerland/GE-SNRCI-7725/2020\|EPI_ISL_548 | ···················· | ···················· | ···················· | ···················· | ···················· | ···················· | |
| I | 2283 hCoV-19/Canada/ON-PHL-20-01311/2020\|EPI_ISL_548914 | ···················· | ···················· | ···················· | ···················· | ···················· | ···················· | |
| I | 2285 hCoV-19/Canada/ON-PHL-20-01314/2020\|EPI_ISL_548917 | ···················· | ···················· | ···················· | ···················· | ···················· | ···················· | |
| I | 2286 hCoV-19/Canada/ON-PHL-20-01315/2020\|EPI_ISL_548918 | ············T······· | ···················· | ···················· | ···················· | ···················· | ···················· | |
| I | 2287 hCoV-19/Canada/ON-PHL-20-01316/2020\|EPI_ISL_548919 | ···················· | ···················· | ···················· | ···················· | ···················· | ···················· | |
| I | 2288 hCoV-19/Canada/ON-PHL-20-01321/2020\|EPI_ISL_548924 | ···················· | ···················· | ···················· | ···················· | ···················· | ···················· | |
| I | 2289 hCoV-19/Canada/ON-PHL-20-01322/2020\|EPI_ISL_548925 | ···················· | ···················· | ···················· | ···················· | ···················· | ···················· | |
| I | 2290 hCoV-19/Canada/ON-PHL-20-01323/2020\|EPI_ISL_548926 | ···················· | ···················· | ···················· | ···················· | ···················· | ···················· | |
| I | 2291 hCoV-19/Canada/ON-PHL-20-01324/2020\|EPI_ISL_548927 | ···················· | ···················· | ···················· | ···················· | ···················· | ···················· | |
| I | 2292 hCoV-19/Canada/ON-PHL-20-01325/2020\|EPI_ISL_548928 | ···················· | ···················· | ···················· | ···················· | ···················· | ···················· | |
| I | 2293 hCoV-19/Canada/ON-PHL-20-01326/2020\|EPI_ISL_548929 | ···················· | ···················· | ···················· | ···················· | ···················· | ···················· | |
| I | 2294 hCoV-19/Canada/ON-PHL-20-01327/2020\|EPI_ISL_548930 | ···················· | ···················· | ···················· | ···················· | ···················· | ···················· | |
| I | 2295 hCoV-19/Canada/ON-PHL-20-01328/2020\|EPI_ISL_548931 | ···················· | ···················· | ···················· | ···················· | ···················· | ···················· | |
| I | 2296 hCoV-19/Canada/ON-PHL-20-90069/2020\|EPI_ISL_548932 | ···················· | ···················· | ···················· | ···················· | ···················· | ···················· | |
| I | 2297 hCoV-19/Canada/ON-PHL-20-90070/2020\|EPI_ISL_548933 | ···················· | ···················· | ···················· | ···················· | ···················· | ···················· | |
| I | 2298 hCoV-19/Canada/ON-PHL-20-90084/2020\|EPI_ISL_548934 | ···················· | ···················· | ···················· | ···················· | ···················· | ···················· | |
| I | 2299 hCoV-19/Canada/ON-PHL-20-90085/2020\|EPI_ISL_548935 | ···················· | ···················· | ···················· | ···················· | ···················· | ···················· | |
| I | 2300 hCoV-19/Canada/ON-PHL-20-90087/2020\|EPI_ISL_548937 | ···················· | ···················· | ···················· | ···················· | ···················· | ···················· | |
| I | 2302 hCoV-19/Canada/ON-PHL-20-90090/2020\|EPI_ISL_548939 | ···················· | ···················· | ···················· | ···················· | ···················· | ···················· | |
| I | 2303 hCoV-19/Canada/ON-PHL-20-90107/2020\|EPI_ISL_548940 | ···················· | ···················· | ···················· | ···················· | ···················· | ···················· | |
| I | 2304 hCoV-19/Canada/ON-PHL-20-90108/2020\|EPI_ISL_548941 | ···················· | ···················· | ···················· | ···················· | ···················· | ···················· | |
| I | 2305 hCoV-19/USA/TX-HMH-4884/2020\|EPI_ISL_544403\|2020-0 | ···················· | ···················· | ···················· | ···················· | ···················· | ···················· | |
| I | 2306 hCoV-19/USA/TX-HMH-4885/2020\|EPI_ISL_544404\|2020-0 | ···················· | ···················· | ···················· | ···················· | ···················· | ···················· | |
| I | 2307 hCoV-19/USA/TX-HMH-489/2020\|EPI_ISL_544406\|2020-04 | ···················· | ···················· | ···················· | ···················· | ···················· | ···················· | |
| I | 2308 hCoV-19/USA/TX-HMH-490/2020\|EPI_ISL_544407\|2020-04 | ···················· | ···················· | ···················· | ···················· | ···················· | ···················· | |
| I | 2309 hCoV-19/USA/TX-HMH-491/2020\|EPI_ISL_544408\|2020-04 | ···················· | ···················· | ···················· | ···················· | ···················· | ···················· | |
| I | 2310 hCoV-19/USA/TX-HMH-492/2020\|EPI_ISL_544409\|2020-04 | ···················· | ···················· | ···················· | ···················· | ···················· | ···················· | |
| I | 2311 hCoV-19/USA/TX-HMH-493/2020\|EPI_ISL_544410\|2020-04 | ···················· | ···················· | ···················· | ···················· | ···················· | ···················· | |
| I | 2313 hCoV-19/USA/TX-HMH-4936/2020\|EPI_ISL_544412\|2020-0 | ···················· | ·············A······ | ···················· | ···················· | ···················· | ···················· | |
| I | 2315 hCoV-19/USA/TX-HMH-494/2020\|EPI_ISL_544415\|2020-04 | ···················· | ···················· | ···················· | ···················· | ···················· | ···················· | |
| I | 2316 hCoV-19/USA/TX-HMH-4940/2020\|EPI_ISL_544416\|2020-0 | ···················· | ···················· | ···················· | ···················· | ···················· | ···················· | |
| I | 2317 hCoV-19/USA/TX-HMH-4945/2020\|EPI_ISL_544421\|2020-0 | ···················· | ···················· | ···················· | ···················· | ···················· | ···················· | |
| I | 2318 hCoV-19/USA/TX-HMH-4947/2020\|EPI_ISL_544422\|2020-0 | ···················· | ···················· | ···················· | ···················· | ···················· | ···················· | |
| I | 2319 hCoV-19/USA/TX-HMH-4948/2020\|EPI_ISL_544423\|2020-0 | ···················· | ···················· | ···················· | ···················· | ···················· | ···················· | |
| I | 2320 hCoV-19/USA/TX-HMH-495/2020\|EPI_ISL_544424\|2020-04 | ···················· | ···················· | ···················· | ···················· | ···················· | ···················· | |
| I | 2321 hCoV-19/USA/TX-HMH-4956/2020\|EPI_ISL_544430\|2020-0 | ···················· | ·············A······ | ···················· | ···················· | ···················· | ···················· | |
| I | 2322 hCoV-19/USA/TX-HMH-5055/2020\|EPI_ISL_544491\|2020-0 | ···················· | ···················· | ···················· | ···················· | ···················· | ···················· | |
| I | 2324 hCoV-19/USA/TX-HMH-5057/2020\|EPI_ISL_544493\|2020-0 | ···················· | ·A·················· | ···················· | ···················· | ···················· | ···················· | |
| I | 2325 hCoV-19/USA/TX-HMH-5060/2020\|EPI_ISL_544495\|2020-0 | ···················· | ···················· | ···················· | ···················· | ···················· | ···················· | |
| I | 2326 hCoV-19/USA/TX-HMH-5061/2020\|EPI_ISL_544496\|2020-0 | ···················· | ···················· | ···················· | ···················· | ···················· | ···················· | |
| I | 2327 hCoV-19/USA/TX-HMH-5062/2020\|EPI_ISL_544497\|2020-0 | ···················· | ···················· | ···················· | ···················· | ···················· | ···················· | |
| I | 2328 hCoV-19/USA/TX-HMH-5063/2020\|EPI_ISL_544498\|2020-0 | ···················· | ···················· | ···················· | ···················· | ···················· | ···················· | |
| I | 2329 hCoV-19/USA/TX-HMH-2054/2020\|EPI_ISL_545090\|2020-0 | ···················· | ···················· | ···················· | ···················· | ···················· | ···················· | |
| I | 2330 hCoV-19/USA/TX-HMH-2064/2020\|EPI_ISL_545098\|2020-0 | ···················· | ···················· | ···················· | ···················· | ···················· | ···················· | |
| I | 2331 hCoV-19/USA/TX-HMH-7951/2020\|EPI_ISL_545734\|2020-0 | ···················· | ···················· | ···················· | ···················· | ···················· | ···················· | |
| I | 2336 hCoV-19/USA/TX-HMH-7957/2020\|EPI_ISL_545739\|2020-0 | ···················· | ····T··············· | ···················· | ···················· | ···················· | ···················· | |
| I | 2338 hCoV-19/USA/TX-HMH-7962/2020\|EPI_ISL_545744\|2020-0 | ···················· | ···················· | ···················· | ···················· | ···················· | ···················· | |
| I | 2339 hCoV-19/USA/TX-HMH-7963/2020\|EPI_ISL_545745\|2020-0 | ···················· | ···················· | ···················· | ···················· | ···················· | ···················· | |
| I | 2341 hCoV-19/USA/TX-HMH-7966/2020\|EPI_ISL_545748\|2020-0 | ···················· | ···················· | ···················· | ···················· | ···················· | ···················· | |
| I | 2343 hCoV-19/USA/TX-HMH-7969/2020\|EPI_ISL_545751\|2020-0 | ···················· | ···················· | ···················· | ···················· | ···················· | ···················· | |
| I | 2344 hCoV-19/USA/TX-HMH-7970/2020\|EPI_ISL_545752\|2020-0 | ···················· | ···················· | ···················· | ···················· | ···················· | ···················· | |
| I | 2348 hCoV-19/USA/TX-HMH-7979/2020\|EPI_ISL_545760\|2020-0 | ···················· | ···················· | ···················· | ···················· | ···················· | ···················· | |
| I | 2349 hCoV-19/USA/TX-HMH-6731/2020\|EPI_ISL_546200\|2020-0 | ···················· | ···················· | ···················· | ···················· | ···················· | ···················· | |
| I | 2350 hCoV-19/USA/TX-HMH-6733/2020\|EPI_ISL_546201\|2020-0 | ···················· | ·············A······ | ···················· | ···················· | ···················· | ···················· | |
| I | 2351 hCoV-19/USA/TX-HMH-6735/2020\|EPI_ISL_546202\|2020-0 | ···················· | ···················· | ···················· | ···················· | ···················· | ···N·········NN····· | 1 |
| I | 2353 hCoV-19/USA/TX-HMH-6740/2020\|EPI_ISL_546207\|2020-0 | ···················· | ···················· | ···················· | ···················· | ···················· | ···················· | |
| I | 2354 hCoV-19/USA/TX-HMH-6741/2020\|EPI_ISL_546208\|2020-0 | ···················· | ···················· | ···················· | ···················· | ···················· | ···················· | |
| I | 2355 hCoV-19/USA/TX-HMH-6742/2020\|EPI_ISL_546209\|2020-0 | ···················· | ···················· | ···················· | ···················· | ···················· | ···················· | |
| I | 2357 hCoV-19/USA/TX-HMH-6746/2020\|EPI_ISL_546211\|2020-0 | ···················· | ···················· | ···················· | ···················· | ···················· | NN·················· | |
| I | 2359 hCoV-19/USA/TX-HMH-6749/2020\|EPI_ISL_546213\|2020-0 | ···················· | ···················· | ···················· | ···················· | ···················· | ···················· | |
| I | 2361 hCoV-19/USA/TX-HMH-6751/2020\|EPI_ISL_546216\|2020-0 | ···················· | ···················· | ···················· | ···················· | ···················· | ···················· | |
| I | 2362 hCoV-19/USA/TX-HMH-6752/2020\|EPI_ISL_546217\|2020-0 | ···················· | ···················· | ···················· | ···················· | ···················· | ···················· | |
| I | 2363 hCoV-19/USA/TX-HMH-6753/2020\|EPI_ISL_546218\|2020-0 | ···················· | ···················· | ···················· | ···················· | ···················· | ···················· | |
| I | 2364 hCoV-19/USA/TX-HMH-6754/2020\|EPI_ISL_546219\|2020-0 | ···················· | ·············A······ | ···················· | ···················· | ···················· | ···················· | |
| I | 2365 hCoV-19/USA/TX-HMH-6755/2020\|EPI_ISL_546220\|2020-0 | ···················· | ···················· | ···················· | ···················· | ···················· | ···················· | |
| I | 2366 hCoV-19/USA/TX-HMH-6756/2020\|EPI_ISL_546221\|2020-0 | ···················· | ···················· | ···················· | ···················· | ···················· | ···················· | |

I 2367 hCoV-19/USA/TX-HMH-6758/2020|EPI_ISL_546222|2020-0 ···················· ·············A······ ···················· ···················· ···················· ····················
I 2369 hCoV-19/USA/TX-HMH-6762/2020|EPI_ISL_546225|2020-0 ···················· ···················· ···················· ···················· ···················· ····················
I 2371 hCoV-19/USA/TX-HMH-6764/2020|EPI_ISL_546227|2020-0 ···················· ·············A······ ···················· ···················· ···················· ····················
I 2373 hCoV-19/USA/TX-HMH-6768/2020|EPI_ISL_546231|2020-0 ···················· ···················· ···················· ···················· ···················· ····················
I 2374 hCoV-19/USA/TX-HMH-6769/2020|EPI_ISL_546232|2020-0 ···················· ···················· ···················· ···················· ···················· ····················
I 2375 hCoV-19/USA/TX-HMH-6770/2020|EPI_ISL_546233|2020-0 ···················· ···················· ···················· ···················· ···················· ····················
I 2376 hCoV-19/USA/TX-HMH-6771/2020|EPI_ISL_546234|2020-0 ···················· ·············A······ ···················· ···················· ···················· ····················
I 2377 hCoV-19/USA/TX-HMH-6774/2020|EPI_ISL_546235|2020-0 ···················· ···················· ···················· ···················· ···················· ····················
I 2378 hCoV-19/USA/TX-HMH-6777/2020|EPI_ISL_546236|2020-0 ···················· ···················· ···················· ···················· ···················· ····················
I 2379 hCoV-19/USA/TX-HMH-6778/2020|EPI_ISL_546237|2020-0 ···················· ···················· ···················· ···················· ···················· ····················
I 2381 hCoV-19/USA/TX-HMH-6784/2020|EPI_ISL_546240|2020-0 ···················· ···················· ···················· ···················· ···················· ····················
I 2382 hCoV-19/USA/TX-HMH-6785/2020|EPI_ISL_546241|2020-0 ···················· ···················· ···················· ···················· ···················· ····················
I 2383 hCoV-19/USA/TX-HMH-6786/2020|EPI_ISL_546242|2020-0 ···················· ···················· ···················· ···················· ···················· ····················
I 2384 hCoV-19/USA/TX-HMH-6787/2020|EPI_ISL_546243|2020-0 ···················· ···················· ···················· ···················· ···················· ····················
I 2385 hCoV-19/USA/TX-HMH-6790/2020|EPI_ISL_546244|2020-0 ···················· ···················· ···················· ···················· ···················· ····················
I 2386 hCoV-19/USA/TX-HMH-6791/2020|EPI_ISL_546245|2020-0 ···················· ···················· ···················· ···················· ···················· ····················
I 2388 hCoV-19/USA/TX-HMH-6793/2020|EPI_ISL_546247|2020-0 ···················· ···················· ···················· ···················· ···················· ····················
I 2389 hCoV-19/USA/TX-HMH-6794/2020|EPI_ISL_546248|2020-0 ···················· ···················· ···················· ···················· ···················· ····················
I 2391 hCoV-19/USA/TX-HMH-6799/2020|EPI_ISL_546251|2020-0 ···················· ···················· ···················· ···················· ···················· ····················
I 2392 hCoV-19/USA/TX-HMH-6800/2020|EPI_ISL_546252|2020-0 ···················· ···················· ···················· ···················· ···················· ····················
I 2393 hCoV-19/USA/TX-HMH-6803/2020|EPI_ISL_546253|2020-0 ···················· ···················· ···················· ···················· ···················· ····················
I 2394 hCoV-19/USA/TX-HMH-6804/2020|EPI_ISL_546254|2020-0 ···················· ···················· ···················· ···················· ···················· ···A················
I 2395 hCoV-19/USA/TX-HMH-6805/2020|EPI_ISL_546255|2020-0 ···················· ···················· ···················· ···················· ···················· ····················
I 2396 hCoV-19/USA/TX-HMH-6806/2020|EPI_ISL_546256|2020-0 ···················· ···················· ···················· ···················· ···················· ····················
I 2397 hCoV-19/USA/TX-HMH-6807/2020|EPI_ISL_546257|2020-0 ···················· ···················· ···················· ···················· ···················· ····················
I 2398 hCoV-19/USA/TX-HMH-6808/2020|EPI_ISL_546258|2020-0 ···················· ···················· ···················· ···················· ···················· ····················
I 2399 hCoV-19/USA/TX-HMH-6809/2020|EPI_ISL_546259|2020-0 ···················· ···················· ···················· ···················· ···················· NN··················
I 2401 hCoV-19/USA/TX-HMH-6815/2020|EPI_ISL_546262|2020-0 ···················· ···················· ···················· ···················· ···················· ····················
I 2403 hCoV-19/USA/TX-HMH-6819/2020|EPI_ISL_546264|2020-0 ···················· ···················· ···················· ···················· ···················· ····················
I 2407 hCoV-19/Netherlands/UT-RIVM-10055/2020|EPI_ISL_547 ···················· ···················· ···················· ···················· ···················· ····················
I 2408 hCoV-19/Netherlands/UT-RIVM-10059/2020|EPI_ISL_547 ···················· ···················· ···················· ···················· ···················· ····················
I 2409 hCoV-19/Netherlands/UT-RIVM-10060/2020|EPI_ISL_547 ···················· ···················· ···················· ···················· ···················· ····················
I 2410 hCoV-19/Netherlands/UT-RIVM-10064/2020|EPI_ISL_547 ···················· ···················· ···················· ···················· ···················· ····················
I 2411 hCoV-19/Netherlands/UT-RIVM-10065/2020|EPI_ISL_547 ···················· ···················· ···················· ···················· ···················· ····················
I 2412 hCoV-19/Netherlands/UT-RIVM-10066/2020|EPI_ISL_547 ···················· ···················· ···················· ···················· ···················· ····················
I 2413 hCoV-19/Netherlands/UT-RIVM-10068/2020|EPI_ISL_547 ···················· ···················· ···················· ···················· ···················· ····················
I 2414 hCoV-19/Netherlands/UT-RIVM-10072/2020|EPI_ISL_547 ···················· ···················· ···················· ···················· ···················· ····················
I 2415 hCoV-19/Netherlands/UT-RIVM-10077/2020|EPI_ISL_547 ···················· ···················· ···················· ···················· ···················· ····················
I 2416 hCoV-19/Netherlands/UT-RIVM-10080/2020|EPI_ISL_547 ···················· ···················· ···················· ···················· ···················· ····················
I 2417 hCoV-19/Netherlands/UT-RIVM-10082/2020|EPI_ISL_547 ···················· ···················· ···················· ···················· ···················· ····················
I 2418 hCoV-19/Netherlands/UT-RIVM-10090/2020|EPI_ISL_547 ···················· ···················· ···················· ···················· ···················· ····················
I 2419 hCoV-19/Netherlands/UT-RIVM-10091/2020|EPI_ISL_547 ···················· ···················· ···················· ···················· ···················· ····················
I 2420 hCoV-19/Netherlands/UT-RIVM-10107/2020|EPI_ISL_547 ···················· ···················· ···················· ···················· ···················· ····················
I 2421 hCoV-19/Netherlands/UT-RIVM-10120/2020|EPI_ISL_547 ···················· ···················· ···················· ···················· ···················· ····················
I 2422 hCoV-19/Netherlands/UT-RIVM-10121/2020|EPI_ISL_547 ···················· ···················· ···················· ···················· ···················· ····················
I 2423 hCoV-19/Netherlands/UT-RIVM-10123/2020|EPI_ISL_547 ···················· ···················· ···················· ···················· ···················· ····················
I 2424 hCoV-19/Netherlands/UT-RIVM-10125/2020|EPI_ISL_547 ···················· ···················· ···················· ···················· ···················· ····················
I 2425 hCoV-19/Netherlands/UT-RIVM-10128/2020|EPI_ISL_547 ···················· ···················· ···················· ···················· ···················· ····················
I 2426 hCoV-19/Netherlands/UT-RIVM-10129/2020|EPI_ISL_547 ···················· ···················· ···················· ···················· ···················· ····················
I 2427 hCoV-19/Netherlands/UT-RIVM-10134/2020|EPI_ISL_547 ···················· ···················· ···················· ···················· ···················· ····················
I 2428 hCoV-19/Netherlands/UT-RIVM-10135/2020|EPI_ISL_547 ···················· ···················· ···················· ···················· ···················· ····················
I 2429 hCoV-19/Netherlands/UT-RIVM-10138/2020|EPI_ISL_547 ···················· ···················· ···················· ···················· ···················· ····················
I 2430 hCoV-19/Netherlands/UT-RIVM-10148/2020|EPI_ISL_547 ···················· ···················· ···················· ···················· ···················· ····················
I 2431 hCoV-19/Netherlands/UT-RIVM-10153/2020|EPI_ISL_547 ···················· ···················· ···················· ···················· ···················· ····················
I 2432 hCoV-19/Netherlands/UT-RIVM-10154/2020|EPI_ISL_547 ···················· ···················· ···················· ···················· ···················· ····················
I 2433 hCoV-19/Netherlands/UT-RIVM-10155/2020|EPI_ISL_547 ···················· ···················· ···················· ···················· ···················· ····················
I 2434 hCoV-19/Netherlands/UT-RIVM-10156/2020|EPI_ISL_547 ···················· ···················· ···················· ···················· ···················· ····················
I 2435 hCoV-19/Netherlands/UT-RIVM-10157/2020|EPI_ISL_547 ···················· ···················· ···················· ···················· ···················· ····················
I 2436 hCoV-19/Netherlands/UT-RIVM-10158/2020|EPI_ISL_547 ···················· ···················· ···················· ···················· ···················· ····················
I 2437 hCoV-19/Netherlands/UT-RIVM-10159/2020|EPI_ISL_547 ···················· ···················· ···················· ···················· ···················· ····················
I 2438 hCoV-19/Netherlands/UT-RIVM-10160/2020|EPI_ISL_547 ···················· ···················· ···················· ···················· ···················· ····················
I 2439 hCoV-19/Netherlands/UT-RIVM-10161/2020|EPI_ISL_547 ···················· ···················· ···················· ···················· ···················· ····················
I 2440 hCoV-19/Netherlands/UT-RIVM-10162/2020|EPI_ISL_547 ···················· ···················· ···················· ···················· ···················· ····················
I 2441 hCoV-19/Netherlands/UT-RIVM-10163/2020|EPI_ISL_547 ···················· ···················· ···················· ···················· ···················· ····················
I 2442 hCoV-19/Netherlands/UT-RIVM-10164/2020|EPI_ISL_547 ···················· ···················· ···················· ···················· ···················· ····················
I 2443 hCoV-19/Netherlands/UT-RIVM-10165/2020|EPI_ISL_547 ···················· ···················· ···················· ···················· ···················· ····················
I 2444 hCoV-19/Netherlands/UT-RIVM-10166/2020|EPI_ISL_547 ···················· ···················· ···················· ···················· ···················· ····················
I 2445 hCoV-19/Netherlands/UT-RIVM-10167/2020|EPI_ISL_547 ···················· ···················· ···················· ···················· ···················· ····················
I 2446 hCoV-19/Netherlands/UT-RIVM-10168/2020|EPI_ISL_547 ···················· ···················· ···················· ···················· ···················· ····················
I 2447 hCoV-19/Netherlands/UT-RIVM-10169/2020|EPI_ISL_547 ···················· ···················· ···················· ···················· ···················· ····················
I 2448 hCoV-19/Netherlands/ZH-RIVM-10050/2020|EPI_ISL_547 ···················· ···················· ···················· ···················· ···················· ····················
I 2449 hCoV-19/Netherlands/ZH-RIVM-10063/2020|EPI_ISL_547 ···················· ···················· ···················· ···················· ···················· ····················
I 2450 hCoV-19/Netherlands/ZH-RIVM-10076/2020|EPI_ISL_547 ···················· ···················· ···················· ···················· ···················· ····················
I 2451 hCoV-19/Netherlands/ZH-RIVM-10079/2020|EPI_ISL_547 ···················· ···················· ···················· ···················· ···················· ····················
I 2452 hCoV-19/Netherlands/ZH-RIVM-10099/2020|EPI_ISL_547 ···················· ···················· ···················· ···················· ···················· ····················
I 2453 hCoV-19/Netherlands/ZH-RIVM-10110/2020|EPI_ISL_547 ···················· ···················· ···················· ···················· ···················· ····················
I 2454 hCoV-19/Netherlands/ZH-RIVM-10116/2020|EPI_ISL_547 ···················· ···················· ···················· ···················· ···················· ····················
I 2455 hCoV-19/Netherlands/ZH-RIVM-10117/2020|EPI_ISL_547 ···················· ···················· ···················· ···················· ···················· ····················
I 2456 hCoV-19/Netherlands/ZH-RIVM-10118/2020|EPI_ISL_547 ···················· ···········A········ ···················· ···················· ···················· ····················
I 2457 hCoV-19/Netherlands/ZH-RIVM-10126/2020|EPI_ISL_547 ···················· ···················· ···················· ···················· ···················· ····················

| | | | | | | | | | |
|---|---|---|---|---|---|---|---|---|---|
| I | 2458 hCoV-19/Netherlands/ZH-RIVM-10127/2020\|EPI_ISL_547 | .................... | .................... | .................... | .................... | .................... | .................... | | |
| I | 2459 hCoV-19/Netherlands/ZH-RIVM-10133/2020\|EPI_ISL_547 | .................... | .................... | .................... | .................... | .................... | .................... | | |
| I | 2460 hCoV-19/Brazil/SP-314/2020\|EPI_ISL_547570\|2020-04- | .................... | .................... | .................... | .................... | .................... | .................... | | |
| I | 2461 hCoV-19/Brazil/SP-341/2020\|EPI_ISL_547571\|2020-06- | .................... | .................... | .................... | .................... | .................... | .................... | | |
| I | 2462 hCoV-19/Brazil/SP-342/2020\|EPI_ISL_547573\|2020-06- | .................... | .................... | .................... | .................... | .................... | .................... | | |
| I | 2463 hCoV-19/Brazil/SP-354/2020\|EPI_ISL_547574\|2020-06- | .................... | .................... | .................... | .................... | .................... | .................... | | |
| I | 2464 hCoV-19/Brazil/SP-356/2020\|EPI_ISL_547575\|2020-06- | .................... | .................... | .................... | .................... | .................... | .................... | | |
| I | 2465 hCoV-19/Brazil/SP-358/2020\|EPI_ISL_547576\|2020-06- | .................... | .................... | .................... | .................... | .................... | .................... | | |
| I | 2466 hCoV-19/Brazil/SP-405/2020\|EPI_ISL_547577\|2020-07- | .................... | .................... | .................... | .................... | .................... | .................... | | |
| I | 2467 hCoV-19/Brazil/SP-406/2020\|EPI_ISL_547578\|2020-07- | .................... | .................... | .................... | .................... | .................... | .................... | | |
| I | 2468 hCoV-19/Brazil/SP-417/2020\|EPI_ISL_547579\|2020-07- | .................... | .................... | .................... | .................... | .................... | .................... | | |
| I | 2469 hCoV-19/Brazil/SP-418/2020\|EPI_ISL_547580\|2020-07- | .................... | .................... | .................... | .................... | .................... | .................... | | |
| I | 2470 hCoV-19/India/UT-IMT-BY60/2020\|EPI_ISL_547584\|2020 | .................... | .A.................. | .................... | .................... | .................... | ...A................ | | |
| I | 2471 hCoV-19/India/HR-IMT-BZ16/2020\|EPI_ISL_547585\|2020 | .................... | .A.................. | .................... | .................... | .................... | ...A................ | | |
| I | 2472 hCoV-19/India/HR-IMT-BZ102/2020\|EPI_ISL_547586\|202 | .................... | .A.................. | .................... | .................... | .................... | ...A................ | | |
| I | 2473 hCoV-19/India/HR-IMT-BZ140/2020\|EPI_ISL_547588\|202 | .................... | .A.................. | .................... | .................... | .................... | ...A................ | | |
| I | 2474 hCoV-19/India/HR-IMT-CB29/2020\|EPI_ISL_547589\|2020 | .................... | .A.................. | .................... | .................... | .................... | ...A................ | | |
| I | 2475 hCoV-19/India/HR-IMT-CB44/2020\|EPI_ISL_547590\|2020 | .................... | .A.................. | .................... | .................... | .................... | ...A................ | | |
| I | 2476 hCoV-19/India/HR-IMT-CB45/2020\|EPI_ISL_547591\|2020 | .................... | .................... | .................... | .................... | .................... | .................... | | |
| I | 2477 hCoV-19/India/HR-IMT-CB130/2020\|EPI_ISL_547592\|202 | .................... | .A.................. | .................... | .................... | .................... | .................... | | |
| I | 2478 hCoV-19/India/HR-IMT-CB193/2020\|EPI_ISL_547593\|202 | .................... | .A.................. | .................... | .................... | .................... | ...A................ | | |
| I | 2479 hCoV-19/Australia/NSW929/2020\|EPI_ISL_547594\|2020- | ........T........... | .................... | .................... | .................... | .................... | .................... | | |
| I | 2480 hCoV-19/Australia/NSW930/2020\|EPI_ISL_547595\|2020- | .................... | .................... | .................... | .................... | .................... | .................... | | |
| I | 2481 hCoV-19/Australia/NSW931/2020\|EPI_ISL_547596\|2020- | .................... | .................... | .................... | .................... | .................... | .................... | | |
| I | 2482 hCoV-19/Australia/NSW932/2020\|EPI_ISL_547597\|2020- | .................... | .................... | .................... | .................... | .................... | .................... | | |
| I | 2483 hCoV-19/Australia/NSW933/2020\|EPI_ISL_547598\|2020- | ........T........... | .................... | .................... | .................... | .................... | .................... | | |
| I | 2484 hCoV-19/Australia/NSW934/2020\|EPI_ISL_547599\|2020- | ........T........... | .................... | .................... | .................... | .................... | .................... | | |
| I | 2485 hCoV-19/New Zealand/20CV0238/2020\|EPI_ISL_548120\|2 | .................... | .A.................. | .................... | .................... | .................... | ...A................ | | |
| I | 2486 hCoV-19/New Zealand/20VR2096/2020\|EPI_ISL_548124\|2 | .................... | .................... | .................... | .................... | .................... | .................... | | |
| I | 2487 hCoV-19/New Zealand/20VR2581/2020\|EPI_ISL_548127\|2 | .................... | .................... | .................... | .................... | .................... | .................... | | |
| I | 2488 hCoV-19/New Zealand/20VR3180/2020\|EPI_ISL_548129\|2 | .................... | .................... | .................... | .................... | .................... | .................... | | |
| I | 2489 hCoV-19/New Zealand/20VR3755/2020\|EPI_ISL_548132\|2 | .................... | .A.................. | .................... | .................... | .................... | .................... | | |
| I | 2490 hCoV-19/New Zealand/20VR3804/2020\|EPI_ISL_548139\|2 | .................... | .................... | .................... | .................... | .................... | .................... | | |
| I | 2491 hCoV-19/New Zealand/20VR3828/2020\|EPI_ISL_548140\|2 | .................... | .................... | .................... | .................... | .................... | .................... | | |
| I | 2492 hCoV-19/New Zealand/20VR3848/2020\|EPI_ISL_548141\|2 | .................... | .................... | .................... | .................... | .................... | .................... | | |
| I | 2493 hCoV-19/New Zealand/20VR3852/2020\|EPI_ISL_548142\|2 | .................... | .................... | .................... | .................... | .................... | .................... | | |
| I | 2494 hCoV-19/New Zealand/20VR3859/2020\|EPI_ISL_548143\|2 | .................... | .................... | .................... | .................... | .................... | .................... | | |
| I | 2495 hCoV-19/Switzerland/FR-SNRCI-7972/2020\|EPI_ISL_548 | .................... | .................... | .................... | .................... | .................... | .................... | | |
| I | 2496 hCoV-19/Switzerland/GE-SNRCI-1227/2020\|EPI_ISL_548 | .................... | ...................A | .................... | .................... | .................... | .................... | 1 | |
| I | 2497 hCoV-19/Switzerland/GE-SNRCI-20/2020\|EPI_ISL_54814 | .................... | .................... | .................... | .................... | .................... | .................... | | |
| I | 2498 hCoV-19/Switzerland/GE-SNRCI-2315/2020\|EPI_ISL_548 | .................... | .................... | .................... | .................... | .................... | .................... | | |
| I | 2499 hCoV-19/Switzerland/GE-SNRCI-2351/2020\|EPI_ISL_548 | .................... | .................... | .................... | .................... | .................... | .................... | | |
| I | 2501 hCoV-19/Switzerland/GE-SNRCI-2480/2020\|EPI_ISL_548 | .................... | .................... | .................... | .................... | .................... | .................... | | |
| I | 2502 hCoV-19/Switzerland/GE-SNRCI-2482/2020\|EPI_ISL_548 | .................... | .................... | .................... | .................... | .................... | .................... | | |
| I | 2503 hCoV-19/Switzerland/GE-SNRCI-2578/2020\|EPI_ISL_548 | .................... | .................... | .................... | .................... | .................... | .................... | | |
| I | 2504 hCoV-19/Switzerland/GE-SNRCI-2616/2020\|EPI_ISL_548 | .................... | .................... | .................... | .................... | .................... | .................... | | |
| I | 2505 hCoV-19/Switzerland/GE-SNRCI-2655/2020\|EPI_ISL_548 | .................... | .................... | .................... | .................... | .................... | .................... | | |
| I | 2506 hCoV-19/Switzerland/GE-SNRCI-2798/2020\|EPI_ISL_548 | .................... | .................... | .................... | .................... | .................... | .................... | | |
| I | 2507 hCoV-19/Switzerland/GE-SNRCI-2825/2020\|EPI_ISL_548 | .................... | .................... | .................... | .................... | .................... | .................... | | |
| I | 2508 hCoV-19/Switzerland/GE-SNRCI-2895/2020\|EPI_ISL_548 | .................... | .................... | .................... | .................... | .................... | .................... | | |
| I | 2509 hCoV-19/Switzerland/GE-SNRCI-2956/2020\|EPI_ISL_548 | .................... | .................... | .................... | .................... | .................... | .................... | | |
| I | 2511 hCoV-19/Switzerland/GE-SNRCI-3187/2020\|EPI_ISL_548 | .................... | .................... | .................... | .................... | .................... | .................... | | |
| I | 2512 hCoV-19/Switzerland/GE-SNRCI-3276/2020\|EPI_ISL_548 | .................... | .A.................. | .................... | .................... | .................... | .................... | | |
| I | 2513 hCoV-19/Switzerland/GE-SNRCI-3403/2020\|EPI_ISL_548 | .................... | .................... | .................... | .................... | .................... | .................... | | |
| I | 2514 hCoV-19/Switzerland/GE-SNRCI-3496/2020\|EPI_ISL_548 | .................... | .................... | .................... | .................... | .................... | .................... | | |
| I | 2515 hCoV-19/Canada/ON-PHL-20-01225/2020\|EPI_ISL_548847 | .................... | .................... | .................... | .................... | .................... | .................... | | |
| I | 2516 hCoV-19/Canada/ON-PHL-20-01226/2020\|EPI_ISL_548848 | .................... | .................... | .................... | .................... | .................... | .................... | | |
| I | 2517 hCoV-19/Canada/ON-PHL-20-01227/2020\|EPI_ISL_548849 | .................... | .................... | .................... | .................... | .................... | .................... | | |
| I | 2518 hCoV-19/Canada/ON-PHL-20-01228/2020\|EPI_ISL_548850 | .................... | .................... | .................... | .................... | NNN................. | .................... | | 1 |
| I | 2520 hCoV-19/Canada/ON-PHL-20-01230/2020\|EPI_ISL_548852 | .................... | .................... | .................... | .................... | .................... | .................... | | |
| I | 2521 hCoV-19/Canada/ON-PHL-20-01231/2020\|EPI_ISL_548853 | .................... | .................... | .................... | .................... | NN.................. | .................... | | |
| I | 2522 hCoV-19/Canada/ON-PHL-20-01232/2020\|EPI_ISL_548854 | .................... | .................... | .................... | .................... | .................... | .................... | | |
| I | 2523 hCoV-19/Canada/ON-PHL-20-01233/2020\|EPI_ISL_548855 | .................... | .................... | .................... | .................... | .................... | .................... | | |
| I | 2525 hCoV-19/Canada/ON-PHL-20-01235/2020\|EPI_ISL_548857 | .................... | ......A............. | .................... | .................... | .................... | .................... | | |
| I | 2526 hCoV-19/Canada/ON-PHL-20-01236/2020\|EPI_ISL_548858 | .................... | .................... | .................... | .................... | .................... | .................... | | |
| I | 2527 hCoV-19/Canada/ON-PHL-20-01237/2020\|EPI_ISL_548859 | .................... | .................... | .................... | .................... | .................... | .................... | | |
| I | 2528 hCoV-19/Canada/ON-PHL-20-01238/2020\|EPI_ISL_548860 | .................... | .................... | .................... | .................... | .................... | .................... | | |
| I | 2529 hCoV-19/Canada/ON-PHL-20-01239/2020\|EPI_ISL_548861 | .................... | .................... | .................... | .................... | .................... | .................... | | |
| I | 2530 hCoV-19/Canada/ON-PHL-20-01240/2020\|EPI_ISL_548862 | .................... | .................... | .................... | .................... | .................... | .................... | | |
| I | 2531 hCoV-19/Canada/ON-PHL-20-01242/2020\|EPI_ISL_548864 | .................... | .................... | .................... | .................... | .................... | .................... | | |
| I | 2532 hCoV-19/Canada/ON-PHL-20-01243/2020\|EPI_ISL_548865 | .................... | .................... | .................... | .................... | .................... | .................... | | |
| I | 2534 hCoV-19/Canada/ON-PHL-20-01246/2020\|EPI_ISL_548868 | .................... | .................... | .................... | .................... | .................... | .................... | | |
| I | 2535 hCoV-19/Canada/ON-PHL-20-01247/2020\|EPI_ISL_548869 | .................... | .................... | .................... | .................... | .................... | .................... | | |
| I | 2536 hCoV-19/Canada/ON-PHL-20-01248/2020\|EPI_ISL_548870 | .................... | .................... | .................... | .................... | .................... | .................... | | |
| I | 2537 hCoV-19/Canada/ON-PHL-20-01249/2020\|EPI_ISL_548871 | .................... | .................... | .................... | .................... | .................... | .................... | | |
| I | 2540 hCoV-19/Canada/ON-PHL-20-01256/2020\|EPI_ISL_548874 | .................... | .................... | .................... | .................... | .................... | .................... | | |
| I | 2542 hCoV-19/Canada/ON-PHL-20-01261/2020\|EPI_ISL_548876 | .................... | .................... | .................... | .................... | .................... | .................... | | |
| I | 2544 hCoV-19/Canada/ON-PHL-20-01264/2020\|EPI_ISL_548878 | .................... | .................... | .................... | .................... | NNN................. | .................... | | 1 |
| I | 2545 hCoV-19/Canada/ON-PHL-20-01265/2020\|EPI_ISL_548879 | .................... | .................... | .................... | .................... | .................... | .................... | | |
| I | 2547 hCoV-19/Canada/ON-PHL-20-01277/2020\|EPI_ISL_548884 | .................... | .................... | .................... | .................... | .................... | .................... | | |

| | | | | | | | | | |
|---|---|---|---|---|---|---|---|---|---|
| I | 2548 | hCoV-19/Canada/ON-PHL-20-01280/2020\|EPI_ISL_548886 | ···················· | ···················· | ···················· | ···················· | ···················· | ···················· | |
| I | 2549 | hCoV-19/Canada/ON-PHL-20-01282/2020\|EPI_ISL_548887 | ···················· | ···················· | ···················· | ···················· | ···················· | ···················· | |
| I | 2550 | hCoV-19/Canada/ON-PHL-20-01283/2020\|EPI_ISL_548888 | ···················· | ···················· | ···················· | ···················· | ···················· | ···················· | |
| I | 2551 | hCoV-19/Canada/ON-PHL-20-01284/2020\|EPI_ISL_548889 | ···················· | ···················· | ···················· | ···················· | ···················· | ···················· | |
| I | 2552 | hCoV-19/Canada/ON-PHL-20-01285/2020\|EPI_ISL_548890 | ···················· | ···················· | ···················· | ···················· | ···················· | ···················· | |
| I | 2553 | hCoV-19/Canada/ON-PHL-20-01286/2020\|EPI_ISL_548891 | ···················· | ···················· | ···················· | ···················· | ···················· | ···················· | |
| I | 2554 | hCoV-19/Canada/ON-PHL-20-01287/2020\|EPI_ISL_548892 | ···················· | ···················· | ···················· | ···················· | ···················· | ···········NN······· | |
| I | 2556 | hCoV-19/Canada/ON-PHL-20-01290/2020\|EPI_ISL_548894 | ···················· | ···················· | ···················· | ···················· | NNN················· | ···················· | 1 |
| I | 2557 | hCoV-19/Canada/ON-PHL-20-01291/2020\|EPI_ISL_548895 | ···················· | ···················· | ···················· | ···················· | ···················· | ···················· | |
| I | 2558 | hCoV-19/Canada/ON-PHL-20-01292/2020\|EPI_ISL_548896 | ···················· | ···················· | ···················· | ···················· | ···················· | ···················· | |
| I | 2560 | hCoV-19/Canada/ON-PHL-20-01295/2020\|EPI_ISL_548899 | ···················· | ···················· | ···················· | ···················· | ···················· | ···················· | |
| I | 2561 | hCoV-19/USA/TX-HMH-4951/2020\|EPI_ISL_544425\|2020-0 | ···················· | ·············A······ | ···················· | ···················· | ···················· | ···················· | |
| I | 2563 | hCoV-19/USA/TX-HMH-4954/2020\|EPI_ISL_544428\|2020-0 | ···················· | ···················· | ···················· | ···················· | ···················· | ···N················ | |
| I | 2564 | hCoV-19/USA/TX-HMH-4955/2020\|EPI_ISL_544429\|2020-0 | ···················· | ···················· | ···················· | ···················· | ···················· | ···················· | |
| I | 2565 | hCoV-19/USA/TX-HMH-4957/2020\|EPI_ISL_544431\|2020-0 | ···················· | ···················· | ···················· | ···················· | ···················· | ···················· | |
| I | 2567 | hCoV-19/USA/TX-HMH-496/2020\|EPI_ISL_544433\|2020-04 | ···················· | ···················· | ···················· | ···················· | ···················· | ···················· | |
| I | 2568 | hCoV-19/USA/TX-HMH-497/2020\|EPI_ISL_544434\|2020-04 | ···················· | ···················· | ···················· | ···················· | ···················· | ···················· | |
| I | 2569 | hCoV-19/USA/TX-HMH-498/2020\|EPI_ISL_544435\|2020-04 | ···················· | ···················· | ···················· | ···················· | ···················· | ···················· | |
| I | 2570 | hCoV-19/USA/TX-HMH-4984/2020\|EPI_ISL_544436\|2020-0 | ···················· | ···················· | ···················· | ···················· | ···················· | N··················· | |
| I | 2571 | hCoV-19/USA/TX-HMH-4988/2020\|EPI_ISL_544439\|2020-0 | ···················· | ·············A······ | ···················· | ···················· | ···················· | ···················· | |
| I | 2572 | hCoV-19/USA/TX-HMH-4989/2020\|EPI_ISL_544440\|2020-0 | ···················· | ···················· | ···················· | ···················· | ···················· | ···················· | |
| I | 2579 | hCoV-19/USA/TX-HMH-5007/2020\|EPI_ISL_544454\|2020-0 | ···················· | ···················· | ···················· | ···················· | ···················· | ···················· | |
| I | 2580 | hCoV-19/USA/TX-HMH-501/2020\|EPI_ISL_544456\|2020-04 | ···················· | ···················· | ···················· | ···················· | ···················· | ···················· | |
| I | 2581 | hCoV-19/USA/TX-HMH-5011/2020\|EPI_ISL_544457\|2020-0 | ···················· | ···················· | ···················· | ···················· | ···················· | ···················· | |
| I | 2584 | hCoV-19/USA/TX-HMH-5014/2020\|EPI_ISL_544460\|2020-0 | ···················· | ·············A······ | ···················· | ···················· | ···················· | ···················· | |
| I | 2585 | hCoV-19/USA/TX-HMH-5015/2020\|EPI_ISL_544461\|2020-0 | ···················· | ···················· | ···················· | ···················· | ···················· | ···················· | |
| I | 2586 | hCoV-19/USA/TX-HMH-5016/2020\|EPI_ISL_544462\|2020-0 | ···················· | ···················· | ···················· | ···················· | ···················· | ···················· | |
| I | 2587 | hCoV-19/USA/TX-HMH-502/2020\|EPI_ISL_544465\|2020-04 | ···················· | ···················· | ···················· | ···················· | ···················· | ···················· | |
| I | 2588 | hCoV-19/USA/TX-HMH-5020/2020\|EPI_ISL_544466\|2020-0 | ···················· | ···················· | ···················· | ···················· | ···················· | ···················· | |
| I | 2591 | hCoV-19/USA/TX-HMH-5028/2020\|EPI_ISL_544469\|2020-0 | ···················· | ···················· | ···················· | ···················· | ···················· | ···················· | |
| I | 2592 | hCoV-19/USA/TX-HMH-5029/2020\|EPI_ISL_544470\|2020-0 | ···················· | ·············A······ | ···················· | ···················· | ···················· | ···················· | |
| I | 2593 | hCoV-19/USA/TX-HMH-503/2020\|EPI_ISL_544471\|2020-04 | ···················· | ···················· | ···················· | ···················· | ···················· | ···················· | |
| I | 2594 | hCoV-19/USA/TX-HMH-5033/2020\|EPI_ISL_544473\|2020-0 | ···················· | ·············A······ | ···················· | ···················· | ···················· | ···················· | |
| I | 2595 | hCoV-19/USA/TX-HMH-5034/2020\|EPI_ISL_544474\|2020-0 | ···················· | ·············A······ | ···················· | ···················· | ···················· | ···················· | |
| I | 2597 | hCoV-19/USA/TX-HMH-5036/2020\|EPI_ISL_544476\|2020-0 | ···················· | ···················· | ···················· | ···················· | ···················· | ···················· | |
| I | 2598 | hCoV-19/USA/TX-HMH-5038/2020\|EPI_ISL_544478\|2020-0 | ···················· | ·············A······ | ···················· | ···················· | ···················· | ···················· | |
| I | 2599 | hCoV-19/USA/TX-HMH-504/2020\|EPI_ISL_544479\|2020-04 | ···················· | ···················· | ···················· | ···················· | ···················· | ···················· | |
| I | 2600 | hCoV-19/USA/TX-HMH-5041/2020\|EPI_ISL_544480\|2020-0 | ···················· | ···················· | ···················· | ···················· | ···················· | ···················· | |
| I | 2601 | hCoV-19/USA/TX-HMH-5042/2020\|EPI_ISL_544481\|2020-0 | ···················· | ·A·················· | ···················· | ···················· | ···················· | ···················· | |
| I | 2602 | hCoV-19/USA/TX-HMH-5045/2020\|EPI_ISL_544482\|2020-0 | ···················· | ···················· | ···················· | ···················· | ···················· | ···················· | |
| I | 2603 | hCoV-19/USA/TX-HMH-5048/2020\|EPI_ISL_544483\|2020-0 | ···················· | ···················· | ···················· | ···················· | ···················· | ···················· | |
| I | 2605 | hCoV-19/USA/TX-HMH-5051/2020\|EPI_ISL_544487\|2020-0 | ···················· | ·A·················· | ···················· | ···················· | ···················· | ···················· | |
| I | 2606 | hCoV-19/USA/TX-HMH-5053/2020\|EPI_ISL_544489\|2020-0 | ···················· | ···················· | ···················· | ···················· | ···················· | ···················· | |
| I | 2607 | hCoV-19/USA/TX-HMH-5069/2020\|EPI_ISL_544501\|2020-0 | ···················· | ···················· | ···················· | ···················· | ···················· | ···················· | |
| I | 2608 | hCoV-19/USA/TX-HMH-5070/2020\|EPI_ISL_544502\|2020-0 | ···················· | ···················· | ···················· | ···················· | ···················· | ···················· | |
| I | 2609 | hCoV-19/USA/TX-HMH-5072/2020\|EPI_ISL_544504\|2020-0 | ···················· | ···················· | ···················· | ···················· | ···················· | ···················· | |
| I | 2610 | hCoV-19/USA/TX-HMH-5074/2020\|EPI_ISL_544505\|2020-0 | ···················· | ···················· | ···················· | ···················· | ···················· | ···················· | |
| I | 2611 | hCoV-19/USA/TX-HMH-5076/2020\|EPI_ISL_544506\|2020-0 | ···················· | ·············A······ | ···················· | ···················· | ···················· | ···················· | |
| I | 2612 | hCoV-19/USA/TX-HMH-5077/2020\|EPI_ISL_544507\|2020-0 | ···················· | ···················· | ···················· | ···················· | ···················· | ···················· | |
| I | 2613 | hCoV-19/USA/TX-HMH-5078/2020\|EPI_ISL_544508\|2020-0 | ···················· | ···················· | ···················· | ···················· | ···················· | ···················· | |
| I | 2614 | hCoV-19/USA/TX-HMH-5082/2020\|EPI_ISL_544511\|2020-0 | ···················· | ···················· | ···················· | ···················· | ···················· | ···················· | |
| I | 2615 | hCoV-19/USA/TX-HMH-5083/2020\|EPI_ISL_544512\|2020-0 | ···················· | ···················· | ···················· | ···················· | ···················· | ···················· | |
| I | 2616 | hCoV-19/USA/TX-HMH-5085/2020\|EPI_ISL_544513\|2020-0 | ···················· | ···················· | ···················· | ···················· | ···················· | ···················· | |
| I | 2617 | hCoV-19/USA/TX-HMH-5086/2020\|EPI_ISL_544514\|2020-0 | ···················· | ·············A······ | ···················· | ···················· | ···················· | ···················· | |
| I | 2619 | hCoV-19/USA/TX-HMH-509/2020\|EPI_ISL_544517\|2020-04 | ···················· | ···················· | ···················· | ···················· | ···················· | ···················· | |
| I | 2620 | hCoV-19/USA/TX-HMH-5091/2020\|EPI_ISL_544518\|2020-0 | ···················· | ·············A······ | ···················· | ···················· | ···················· | ···················· | |
| I | 2621 | hCoV-19/USA/TX-HMH-5092/2020\|EPI_ISL_544519\|2020-0 | ···················· | ···················· | ···················· | ···················· | ···················· | ···················· | |
| I | 2622 | hCoV-19/USA/TX-HMH-5094/2020\|EPI_ISL_544520\|2020-0 | ···················· | ···················· | ···················· | ···················· | ···················· | ···················· | |
| I | 2624 | hCoV-19/USA/TX-HMH-5096/2020\|EPI_ISL_544522\|2020-0 | ···················· | ···················· | ···················· | ···················· | ···················· | ···················· | |
| I | 2625 | hCoV-19/USA/TX-HMH-5098/2020\|EPI_ISL_544523\|2020-0 | ···················· | ···················· | ···················· | ···················· | ···················· | ···················· | |
| I | 2627 | hCoV-19/USA/TX-HMH-7975/2020\|EPI_ISL_545757\|2020-0 | ···················· | ···················· | ···················· | ···················· | ···················· | ···················· | |
| I | 2628 | hCoV-19/USA/TX-HMH-7977/2020\|EPI_ISL_545758\|2020-0 | ···················· | ···················· | ···················· | ···················· | ···················· | ···················· | |
| I | 2629 | hCoV-19/USA/TX-HMH-7978/2020\|EPI_ISL_545759\|2020-0 | ···················· | ·A·················· | ···················· | ···················· | ···················· | ···················· | |
| I | 2630 | hCoV-19/USA/TX-HMH-7980/2020\|EPI_ISL_545761\|2020-0 | ···················· | ···················· | ···················· | ···················· | ···················· | N··················· | |
| I | 2634 | hCoV-19/USA/TX-HMH-7984/2020\|EPI_ISL_545765\|2020-0 | ···················· | ···················· | ···················· | ···················· | ···················· | ···················· | |
| I | 2635 | hCoV-19/USA/TX-HMH-7985/2020\|EPI_ISL_545766\|2020-0 | ···················· | ·A·················· | ···················· | ···················· | ···················· | ···················· | |
| I | 2636 | hCoV-19/USA/TX-HMH-7986/2020\|EPI_ISL_545767\|2020-0 | ···················· | ···················· | ···················· | ···················· | ···················· | ···················· | |
| I | 2637 | hCoV-19/USA/TX-HMH-7989/2020\|EPI_ISL_545769\|2020-0 | ···················· | ···················· | ···················· | ···················· | ···················· | ···················· | |
| I | 2638 | hCoV-19/USA/TX-HMH-799/2020\|EPI_ISL_545770\|2020-04 | ···················· | ···················· | ···················· | ···················· | ···················· | ···················· | |
| I | 2639 | hCoV-19/USA/TX-HMH-7990/2020\|EPI_ISL_545771\|2020-0 | ···················· | ···················· | ···················· | ···················· | ···················· | ···················· | |
| I | 2640 | hCoV-19/USA/TX-HMH-7991/2020\|EPI_ISL_545772\|2020-0 | ···················· | ···················· | ···················· | ···················· | ···················· | ···················· | |
| I | 2641 | hCoV-19/USA/TX-HMH-7995/2020\|EPI_ISL_545776\|2020-0 | ···················· | ···················· | ···················· | ···················· | ···················· | ···················· | |
| I | 2642 | hCoV-19/USA/TX-HMH-7997/2020\|EPI_ISL_545777\|2020-0 | ···················· | ···················· | ···················· | ···················· | ···················· | ···················· | |
| I | 2643 | hCoV-19/USA/TX-HMH-7998/2020\|EPI_ISL_545778\|2020-0 | ···················· | ··············C····· | ···········T········ | ···················· | ···················· | ···················· | |
| I | 2645 | hCoV-19/USA/TX-HMH-800/2020\|EPI_ISL_545780\|2020-04 | ···················· | ···················· | ···················· | ···················· | ···················· | ···················· | |
| I | 2646 | hCoV-19/USA/TX-HMH-8004/2020\|EPI_ISL_545782\|2020-0 | ···················· | ···················· | ···················· | ···················· | ···················· | ···················· | |
| I | 2647 | hCoV-19/USA/TX-HMH-8005/2020\|EPI_ISL_545783\|2020-0 | ···················· | ·············A······ | ···················· | ···················· | ···················· | ···················· | |
| I | 2648 | hCoV-19/USA/TX-HMH-8006/2020\|EPI_ISL_545784\|2020-0 | ···················· | ···················· | ···················· | ···················· | ···················· | ···················· | |
| I | 2649 | hCoV-19/USA/TX-HMH-8007/2020\|EPI_ISL_545785\|2020-0 | ···················· | ···················· | ···················· | ···················· | ···················· | ···················· | |
| I | 2650 | hCoV-19/USA/TX-HMH-8008/2020\|EPI_ISL_545786\|2020-0 | ···················· | ···················· | ···················· | ···················· | ···················· | ···················· | |

| | | | | | | | | | |
|---|---|---|---|---|---|---|---|---|---|
| I | 2651 | hCoV-19/USA/TX-HMH-8009/2020\|EPI_ISL_545787\|2020-0 | ···················· | ··············C····· | ··········T········· | ···················· | ··············N····· | ···················· | |
| I | 2652 | hCoV-19/USA/TX-HMH-801/2020\|EPI_ISL_545788\|2020-04 | ···················· | ···················· | ···················· | ···················· | ···················· | ···················· | |
| I | 2653 | hCoV-19/USA/TX-HMH-8010/2020\|EPI_ISL_545789\|2020-0 | ···················· | ···················· | ···················· | ···················· | ···················· | ···················· | |
| I | 2654 | hCoV-19/USA/TX-HMH-8011/2020\|EPI_ISL_545790\|2020-0 | ···················· | ···················· | ···················· | ···················· | ···················· | ···················· | |
| I | 2655 | hCoV-19/USA/TX-HMH-8012/2020\|EPI_ISL_545791\|2020-0 | ···················· | ···················· | ···················· | ···················· | ···················· | ···················· | |
| I | 2656 | hCoV-19/USA/TX-HMH-8016/2020\|EPI_ISL_545793\|2020-0 | ···················· | ···················· | ···················· | ···················· | ···················· | ···················· | |
| I | 2657 | hCoV-19/USA/TX-HMH-8017/2020\|EPI_ISL_545794\|2020-0 | ·T·················· | ···················· | ···················· | ···················· | ···················· | ···················· | |
| I | 2659 | hCoV-19/USA/TX-HMH-8020/2020\|EPI_ISL_545796\|2020-0 | ···················· | ···················· | ···················· | ···················· | ···················· | ···················· | |
| I | 2660 | hCoV-19/USA/TX-HMH-8024/2020\|EPI_ISL_545797\|2020-0 | ···················· | ···················· | ···················· | ···················· | ···················· | ···················· | |
| I | 2661 | hCoV-19/USA/TX-HMH-8026/2020\|EPI_ISL_545798\|2020-0 | ···················· | ·A·················· | ···················· | ···················· | ···················· | ···················· | |
| I | 2663 | hCoV-19/USA/TX-HMH-8029/2020\|EPI_ISL_545800\|2020-0 | ···················· | ···················· | ···················· | ···················· | ···················· | ···················· | |
| I | 2664 | hCoV-19/USA/TX-HMH-803/2020\|EPI_ISL_545801\|2020-04 | ···················· | ···················· | ···················· | ···················· | ···················· | ···················· | |
| I | 2665 | hCoV-19/USA/TX-HMH-8030/2020\|EPI_ISL_545802\|2020-0 | ···················· | ···················· | ···················· | ···················· | ···················· | ···················· | |
| I | 2666 | hCoV-19/USA/TX-HMH-8031/2020\|EPI_ISL_545803\|2020-0 | ···················· | ···················· | ···················· | ···················· | ···················· | ···················· | |
| I | 2667 | hCoV-19/USA/TX-HMH-8032/2020\|EPI_ISL_545804\|2020-0 | ···················· | ···················· | ···················· | ···················· | ···················· | ···················· | |
| I | 2668 | hCoV-19/USA/TX-HMH-8033/2020\|EPI_ISL_545805\|2020-0 | ···················· | ···················· | ···················· | ···················· | ···················· | ···················· | |
| I | 2669 | hCoV-19/USA/TX-HMH-8037/2020\|EPI_ISL_545807\|2020-0 | ···················· | ···················· | ···················· | ···················· | ···················· | ···················· | |
| I | 2671 | hCoV-19/USA/TX-HMH-8039/2020\|EPI_ISL_545809\|2020-0 | ···················· | ···················· | ···················· | ···················· | ···················· | ···················· | |
| I | 2672 | hCoV-19/USA/TX-HMH-804/2020\|EPI_ISL_545810\|2020-04 | ···················· | ···················· | ···················· | ···················· | ···················· | NN·················· | |
| I | 2673 | hCoV-19/USA/TX-HMH-8040/2020\|EPI_ISL_545811\|2020-0 | ···················· | ···················· | ···················· | ···················· | ···················· | ···················· | |
| I | 2674 | hCoV-19/USA/TX-HMH-8041/2020\|EPI_ISL_545812\|2020-0 | ···················· | ···················· | ···················· | ···················· | ···················· | ···················· | |
| I | 2677 | hCoV-19/USA/TX-HMH-8049/2020\|EPI_ISL_545816\|2020-0 | ···················· | ···················· | ···················· | ···················· | ···················· | ···················· | |
| I | 2678 | hCoV-19/USA/TX-HMH-805/2020\|EPI_ISL_545817\|2020-04 | ···················· | ···················· | ···················· | ···················· | ···················· | ···················· | |
| I | 2679 | hCoV-19/USA/TX-HMH-8050/2020\|EPI_ISL_545818\|2020-0 | ···················· | ···················· | ···················· | ···················· | ···················· | NN·················· | |
| I | 2680 | hCoV-19/USA/TX-HMH-8051/2020\|EPI_ISL_545819\|2020-0 | ···················· | ·············A······ | ···················· | ···················· | ···················· | ···················· | |
| I | 2681 | hCoV-19/USA/TX-HMH-8053/2020\|EPI_ISL_545820\|2020-0 | ···················· | ···················· | ···················· | ···················· | ···················· | ···················· | |
| I | 2682 | hCoV-19/USA/TX-HMH-8055/2020\|EPI_ISL_545822\|2020-0 | ···················· | ···················· | ···················· | ···················· | ···················· | ···················· | |
| I | 2683 | hCoV-19/USA/TX-HMH-8056/2020\|EPI_ISL_545823\|2020-0 | ···················· | ···················· | ···················· | ···················· | ···················· | ···················· | |
| I | 2684 | hCoV-19/USA/TX-HMH-8058/2020\|EPI_ISL_545824\|2020-0 | ···················· | ···················· | ···················· | ···················· | ··N··········NN····· | NN·····N············ | 1 |
| I | 2685 | hCoV-19/USA/TX-HMH-8059/2020\|EPI_ISL_545825\|2020-0 | ···················· | ···················· | ···················· | ···················· | ···················· | ···················· | |
| I | 2687 | hCoV-19/USA/TX-HMH-8061/2020\|EPI_ISL_545828\|2020-0 | ···················· | ···················· | ···················· | ···················· | ···················· | ···················· | |
| I | 2689 | hCoV-19/USA/TX-HMH-8064/2020\|EPI_ISL_545830\|2020-0 | ···················· | ·A·················· | ···················· | ···················· | ···················· | ···················· | |
| I | 2690 | hCoV-19/USA/TX-HMH-8066/2020\|EPI_ISL_545831\|2020-0 | ···················· | ·············A······ | ···················· | ···················· | ···················· | ···················· | |
| I | 2691 | hCoV-19/USA/TX-HMH-8067/2020\|EPI_ISL_545832\|2020-0 | ···················· | ···················· | ···················· | ···················· | ···················· | ···················· | |
| I | 2692 | hCoV-19/USA/TX-HMH-8068/2020\|EPI_ISL_545833\|2020-0 | ···················· | ·············A······ | ···················· | ···················· | ···················· | ···················· | |
| I | 2693 | hCoV-19/USA/TX-HMH-8069/2020\|EPI_ISL_545834\|2020-0 | ···················· | ···················· | ···················· | ···················· | ···················· | ···················· | |
| I | 2694 | hCoV-19/USA/TX-HMH-807/2020\|EPI_ISL_545835\|2020-04 | ···················· | ···················· | ···················· | ···················· | ···················· | ···················· | |
| I | 2696 | hCoV-19/USA/TX-HMH-8071/2020\|EPI_ISL_545837\|2020-0 | ···················· | ···················· | ···················· | ···················· | ···················· | ···················· | |
| I | 2697 | hCoV-19/USA/TX-HMH-8072/2020\|EPI_ISL_545838\|2020-0 | ···················· | ···················· | ···················· | ···················· | ···················· | ···················· | |
| I | 2698 | hCoV-19/USA/TX-HMH-8073/2020\|EPI_ISL_545839\|2020-0 | ···················· | ···················· | ···················· | ···················· | ···················· | ···················· | |
| I | 2699 | hCoV-19/USA/TX-HMH-8074/2020\|EPI_ISL_545840\|2020-0 | ···················· | ···················· | ···················· | ···················· | ···················· | ···················· | |
| I | 2700 | hCoV-19/USA/TX-HMH-8076/2020\|EPI_ISL_545841\|2020-0 | ···················· | ···················· | ···················· | ···················· | ···················· | ···················· | |
| I | 2701 | hCoV-19/USA/TX-HMH-8077/2020\|EPI_ISL_545842\|2020-0 | ···················· | ···················· | ···················· | ···················· | ···················· | ···················· | |
| I | 2702 | hCoV-19/USA/TX-HMH-8078/2020\|EPI_ISL_545843\|2020-0 | ···················· | ···················· | ···················· | ···················· | ···················· | ···················· | |
| I | 2703 | hCoV-19/USA/TX-HMH-810/2020\|EPI_ISL_545844\|2020-04 | ···················· | ···················· | ···················· | ···················· | ···················· | ···················· | |
| I | 2704 | hCoV-19/USA/TX-HMH-812/2020\|EPI_ISL_545846\|2020-04 | ···················· | ···················· | ···················· | ···················· | ···················· | ···················· | |
| I | 2705 | hCoV-19/USA/TX-HMH-813/2020\|EPI_ISL_545847\|2020-04 | ···················· | ···················· | ···················· | ···················· | ···················· | ···················· | |
| I | 2706 | hCoV-19/USA/TX-HMH-814/2020\|EPI_ISL_545848\|2020-04 | ···················· | ···················· | ···················· | ···················· | ···················· | ···················· | |
| I | 2707 | hCoV-19/USA/TX-HMH-815/2020\|EPI_ISL_545849\|2020-04 | ···················· | ···················· | ···················· | ···················· | ···················· | ···················· | |
| I | 2708 | hCoV-19/USA/TX-HMH-816/2020\|EPI_ISL_545850\|2020-04 | ···················· | ···················· | ···················· | ···················· | ···················· | ···················· | |
| I | 2709 | hCoV-19/USA/TX-HMH-819/2020\|EPI_ISL_545852\|2020-04 | ···················· | ···················· | ···················· | ···················· | ···················· | ···················· | |
| I | 2710 | hCoV-19/USA/TX-HMH-822/2020\|EPI_ISL_545854\|2020-04 | ···················· | ···················· | ···················· | ···················· | ···················· | ···················· | |
| I | 2711 | hCoV-19/USA/TX-HMH-6829/2020\|EPI_ISL_546272\|2020-0 | ···················· | ···················· | ···················· | ···················· | ···················· | ···················· | |
| I | 2715 | hCoV-19/USA/TX-HMH-684/2020\|EPI_ISL_546280\|2020-04 | ···················· | ···················· | ···················· | ···················· | NNN················· | ···················· | 1 |
| I | 2719 | hCoV-19/USA/TX-HMH-685/2020\|EPI_ISL_546288\|2020-04 | ···················· | ···················· | ···················· | ···················· | ···················· | ···················· | |
| I | 2720 | hCoV-19/USA/TX-HMH-6851/2020\|EPI_ISL_546290\|2020-0 | ···················· | ···················· | ···················· | ···················· | ···················· | ···················· | |
| I | 2724 | hCoV-19/USA/TX-HMH-6857/2020\|EPI_ISL_546294\|2020-0 | ···················· | ···················· | ···················· | ···················· | ···················· | ···················· | |
| I | 2725 | hCoV-19/USA/TX-HMH-686/2020\|EPI_ISL_546295\|2020-04 | ···················· | ···················· | ···················· | ···················· | ···················· | ···················· | |
| I | 2726 | hCoV-19/USA/TX-HMH-6860/2020\|EPI_ISL_546296\|2020-0 | ···················· | ···················· | ···················· | ···················· | ··············N····· | ···················· | |
| I | 2727 | hCoV-19/USA/TX-HMH-6861/2020\|EPI_ISL_546297\|2020-0 | ···················· | ···················· | ···················· | ···················· | ···················· | ···················· | |
| I | 2728 | hCoV-19/USA/TX-HMH-6863/2020\|EPI_ISL_546299\|2020-0 | ···················· | ···················· | ···················· | ···················· | ···················· | ···················· | |
| I | 2729 | hCoV-19/Netherlands/NH-RIVM-10102/2020\|EPI_ISL_547 | ···················· | ···················· | ···················· | ···················· | ···················· | ···················· | |
| I | 2730 | hCoV-19/Netherlands/NH-RIVM-10103/2020\|EPI_ISL_547 | ···················· | ···················· | ···················· | ···················· | ···················· | ···················· | |
| I | 2731 | hCoV-19/Netherlands/NH-RIVM-10104/2020\|EPI_ISL_547 | ···················· | ···················· | ···················· | ···················· | ···················· | ···················· | |
| I | 2732 | hCoV-19/Netherlands/NH-RIVM-10108/2020\|EPI_ISL_547 | ···················· | ···················· | ···················· | ···················· | ···················· | ···················· | |
| I | 2733 | hCoV-19/Netherlands/NH-RIVM-10109/2020\|EPI_ISL_547 | ···················· | ···················· | ···················· | ···················· | ···················· | ···················· | |
| I | 2734 | hCoV-19/Netherlands/NH-RIVM-10137/2020\|EPI_ISL_547 | ···················· | ···················· | ···················· | ···················· | ···················· | ···················· | |
| I | 2735 | hCoV-19/Netherlands/NH-RIVM-10139/2020\|EPI_ISL_547 | ···················· | ···················· | ···················· | ···················· | ···················· | ···················· | |
| I | 2736 | hCoV-19/Netherlands/NH-RIVM-10140/2020\|EPI_ISL_547 | ···················· | ···················· | ···················· | ···················· | ···················· | ···················· | |
| I | 2737 | hCoV-19/Netherlands/NH-RIVM-10141/2020\|EPI_ISL_547 | ···················· | ···················· | ···················· | ···················· | ···················· | ···················· | |
| I | 2738 | hCoV-19/Netherlands/NH-RIVM-10144/2020\|EPI_ISL_547 | ···················· | ···················· | ···················· | ···················· | ···················· | ···················· | |
| I | 2739 | hCoV-19/Netherlands/OV-RIVM-10136/2020\|EPI_ISL_547 | ···················· | ···················· | ···················· | ···················· | ···················· | ···················· | |
| I | 2740 | hCoV-19/Netherlands/OV-RIVM-10145/2020\|EPI_ISL_547 | ···················· | ···················· | ···················· | ···················· | ···················· | ···················· | |
| I | 2741 | hCoV-19/Netherlands/OV-RIVM-10149/2020\|EPI_ISL_547 | ···················· | ···················· | ···················· | ···················· | ···················· | ···················· | |
| I | 2742 | hCoV-19/Netherlands/OV-RIVM-10152/2020\|EPI_ISL_547 | ···················· | ···················· | ···················· | ···················· | ···················· | ···················· | |
| I | 2743 | hCoV-19/Netherlands/UT-RIVM-10046/2020\|EPI_ISL_547 | ···················· | ···················· | ···················· | ···················· | ···················· | ···················· | |
| I | 2744 | hCoV-19/Netherlands/UT-RIVM-10054/2020\|EPI_ISL_547 | ···················· | ···················· | ···················· | ···················· | ···················· | ···················· | |
| I | 2745 | hCoV-19/New Zealand/20CV0108/2020\|EPI_ISL_548000\|2 | ···················· | ···················· | ···················· | ···················· | ···················· | ···················· | |
| I | 2746 | hCoV-19/New Zealand/20CV0116/2020\|EPI_ISL_548002\|2 | ···················· | ···················· | ···················· | ···················· | ···················· | ···················· | |
| I | 2747 | hCoV-19/New Zealand/20CV0118/2020\|EPI_ISL_548004\|2 | ···················· | ···················· | ···················· | ···················· | ···················· | ···················· | |

```
I   2748 hCoV-19/New Zealand/20CV0119/2020|EPI_ISL_548005|2   ····················  ····················  ····················  ····················  ····················  ····················
I   2749 hCoV-19/New Zealand/20CV0120/2020|EPI_ISL_548006|2   ····················  ····················  ····················  ····················  ····················  ····················
I   2750 hCoV-19/New Zealand/20CV0122/2020|EPI_ISL_548008|2   ····················  ····················  ····················  ····················  ····················  ····················
I   2751 hCoV-19/New Zealand/20CV0123/2020|EPI_ISL_548009|2   ····················  ····················  ····················  ····················  ····················  ····················
I   2752 hCoV-19/New Zealand/20CV0124/2020|EPI_ISL_548010|2   ····················  ····················  ····················  ····················  ····················  ····················
I   2753 hCoV-19/New Zealand/20CV0125/2020|EPI_ISL_548011|2   ····················  ····················  ····················  ····················  ····················  ····················
I   2754 hCoV-19/New Zealand/20CV0127/2020|EPI_ISL_548013|2   ····················  ····················  ····················  ····················  ····················  ····················
I   2755 hCoV-19/New Zealand/20CV0128/2020|EPI_ISL_548014|2   ····················  ····················  ····················  ····················  ····················  ····················
I   2756 hCoV-19/New Zealand/20CV0129/2020|EPI_ISL_548015|2   ····················  ····················  ····················  ····················  ····················  ····················
I   2757 hCoV-19/New Zealand/20CV0130/2020|EPI_ISL_548016|2   ····················  ····················  ····················  ····················  ····················  ····················
I   2758 hCoV-19/New Zealand/20CV0133/2020|EPI_ISL_548017|2   ····················  ····················  ····················  ····················  ····················  ····················
I   2759 hCoV-19/New Zealand/20CV0145/2020|EPI_ISL_548023|2   ····················  ····················  ····················  ····················  ····················  ····················
I   2760 hCoV-19/New Zealand/20CV0146/2020|EPI_ISL_548024|2   ····················  ····················  ····················  ····················  ····················  ····················
I   2761 hCoV-19/New Zealand/20CV0183/2020|EPI_ISL_548050|2   ····················  ····················  ····················  ····················  ····················  ····················
I   2762 hCoV-19/New Zealand/20CV0191/2020|EPI_ISL_548057|2   ····················  ····················  ····················  ····················  ····················  ····················
I   2763 hCoV-19/New Zealand/20CV0217/2020|EPI_ISL_548064|2   ····················  ····················  ····················  ····················  ····················  ····················
I   2764 hCoV-19/New Zealand/20CV0220/2020|EPI_ISL_548067|2   ····················  ····················  ····················  ····················  ····················  ····················
I   2765 hCoV-19/New Zealand/20CV0230/2020|EPI_ISL_548069|2   ····················  ····················  ····················  ····················  ····················  ····················
I   2766 hCoV-19/New Zealand/20CV0241/2020|EPI_ISL_548075|2   ····················  ····················  ····················  ····················  ····················  ····················
I   2767 hCoV-19/New Zealand/20CV0246/2020|EPI_ISL_548077|2   ····················  ····················  ····················  ····················  ····················  ····················
I   2768 hCoV-19/New Zealand/20CV0254/2020|EPI_ISL_548081|2   ····················  ····················  ····················  ····················  ····················  ····················
I   2769 hCoV-19/New Zealand/20CV0256/2020|EPI_ISL_548082|2   ····················  ····················  ····················  ····················  ····················  ····················
I   2770 hCoV-19/Canada/ON-PHL-20-01178/2020|EPI_ISL_548800   ····················  ····················  ····················  ····················  ····················  ····················
I   2771 hCoV-19/Canada/ON-PHL-20-01179/2020|EPI_ISL_548801   ····················  ····················  ····················  ····················  ····················  ····················
I   2772 hCoV-19/Canada/ON-PHL-20-01180/2020|EPI_ISL_548802   ····················  ····················  ····················  ····················  ····················  ····················
I   2773 hCoV-19/Canada/ON-PHL-20-01181/2020|EPI_ISL_548803   ····················  ····················  ····················  ····················  ····················  ····················
I   2774 hCoV-19/Canada/ON-PHL-20-01182/2020|EPI_ISL_548804   ············T·······  ····················  ····················  ····················  ····················  ····················
I   2775 hCoV-19/Canada/ON-PHL-20-01183/2020|EPI_ISL_548805   ····················  ····················  ····················  ····················  ····················  ····················
I   2776 hCoV-19/Canada/ON-PHL-20-01184/2020|EPI_ISL_548806   ····················  ····················  ····················  ····················  ····················  ····················
I   2777 hCoV-19/Canada/ON-PHL-20-01185/2020|EPI_ISL_548807   ············T·······  ····················  ····················  ····················  ····················  ····················
I   2778 hCoV-19/Canada/ON-PHL-20-01186/2020|EPI_ISL_548808   ····················  ····················  ····················  ····················  ····················  ····················
I   2779 hCoV-19/Canada/ON-PHL-20-01187/2020|EPI_ISL_548809   ····················  ····················  ····················  ····················  ····················  ····················
I   2780 hCoV-19/Canada/ON-PHL-20-01188/2020|EPI_ISL_548810   ····················  ····················  ····················  ····················  ····················  ····················
I   2781 hCoV-19/Canada/ON-PHL-20-01189/2020|EPI_ISL_548811   ····················  ····················  ····················  ····················  ····················  ····················
I   2782 hCoV-19/Canada/ON-PHL-20-01190/2020|EPI_ISL_548812   ····················  ····················  ····················  ····················  ····················  ····················
I   2783 hCoV-19/Canada/ON-PHL-20-01191/2020|EPI_ISL_548813   ····················  ····················  ····················  ····················  ····················  ····················
I   2784 hCoV-19/Canada/ON-PHL-20-01192/2020|EPI_ISL_548814   ····················  ····················  ····················  ····················  ····················  ····················
I   2785 hCoV-19/Canada/ON-PHL-20-01193/2020|EPI_ISL_548815   ····················  ····················  ····················  ····················  ····················  ····················
I   2786 hCoV-19/Canada/ON-PHL-20-01194/2020|EPI_ISL_548816   ····················  ····················  ····················  ····················  N···················  ····················
I   2787 hCoV-19/Canada/ON-PHL-20-01195/2020|EPI_ISL_548817   ····················  ····················  ····················  ····················  ····················  ····················
I   2788 hCoV-19/Canada/ON-PHL-20-01196/2020|EPI_ISL_548818   ····················  ····················  ····················  ····················  ····················  ····················
I   2789 hCoV-19/Canada/ON-PHL-20-01197/2020|EPI_ISL_548819   ····················  ····················  ····················  ····················  ····················  ····················
I   2790 hCoV-19/Canada/ON-PHL-20-01198/2020|EPI_ISL_548820   ····················  ····················  ····················  ····················  ····················  ····················
I   2791 hCoV-19/Canada/ON-PHL-20-01199/2020|EPI_ISL_548821   ····················  ····················  ····················  ····················  ····················  ····················
I   2792 hCoV-19/Canada/ON-PHL-20-01200/2020|EPI_ISL_548822   ····················  ····················  ····················  ····················  ····················  ····················
I   2793 hCoV-19/Canada/ON-PHL-20-01201/2020|EPI_ISL_548823   ····················  ····················  ····················  ····················  ····················  ····················
I   2794 hCoV-19/Canada/ON-PHL-20-01202/2020|EPI_ISL_548824   ····················  ····················  ····················  ····················  ····················  ····················
I   2795 hCoV-19/Canada/ON-PHL-20-01203/2020|EPI_ISL_548825   ····················  ······A·············  ····················  ····················  ····················  ····················
I   2796 hCoV-19/Canada/ON-PHL-20-01204/2020|EPI_ISL_548826   ····················  ····················  ····················  ····················  ····················  ····················
I   2797 hCoV-19/Canada/ON-PHL-20-01205/2020|EPI_ISL_548827   ····················  ····················  ····················  ····················  ····················  ····················
I   2798 hCoV-19/Canada/ON-PHL-20-01206/2020|EPI_ISL_548828   ····················  ····················  ····················  ····················  ····················  ····················
I   2799 hCoV-19/Canada/ON-PHL-20-01207/2020|EPI_ISL_548829   ····················  ······A·············  ····················  ····················  ····················  ····················
I   2800 hCoV-19/Canada/ON-PHL-20-01208/2020|EPI_ISL_548830   ····················  ····················  ····················  ····················  ····················  ····················
I   2801 hCoV-19/Canada/ON-PHL-20-01209/2020|EPI_ISL_548831   ····················  ····················  ····················  ····················  ····················  ····················
I   2802 hCoV-19/Canada/ON-PHL-20-01210/2020|EPI_ISL_548832   ····················  ····················  ····················  ····················  ····················  ····················
I   2803 hCoV-19/Canada/ON-PHL-20-01211/2020|EPI_ISL_548833   ····················  ····················  ····················  ····················  ····················  ····················
I   2804 hCoV-19/Canada/ON-PHL-20-01212/2020|EPI_ISL_548834   ····················  ····················  ····················  ····················  ····················  ····················
I   2805 hCoV-19/Canada/ON-PHL-20-01213/2020|EPI_ISL_548835   ····················  ····················  ····················  ····················  ····················  ····················
I   2806 hCoV-19/Canada/ON-PHL-20-01214/2020|EPI_ISL_548836   ····················  ····················  ····················  ····················  ····················  ····················
I   2807 hCoV-19/Canada/ON-PHL-20-01215/2020|EPI_ISL_548837   ····················  ····················  ····················  ····················  ····················  ····················
I   2808 hCoV-19/Canada/ON-PHL-20-01216/2020|EPI_ISL_548838   ····················  ····················  ····················  ····················  ····················  ····················
I   2809 hCoV-19/Canada/ON-PHL-20-01217/2020|EPI_ISL_548839   ····················  ····················  ····················  ····················  ····················  ····················
I   2810 hCoV-19/Canada/ON-PHL-20-01218/2020|EPI_ISL_548840   ····················  ····················  ····················  ····················  ····················  ····················
I   2811 hCoV-19/Canada/ON-PHL-20-01219/2020|EPI_ISL_548841   ····················  ····················  ····················  ····················  ····················  ····················
I   2812 hCoV-19/Canada/ON-PHL-20-01220/2020|EPI_ISL_548842   ····················  ······A·············  ····················  ····················  ····················  ····················
I   2813 hCoV-19/Canada/ON-PHL-20-01221/2020|EPI_ISL_548843   ····················  ····················  ····················  ····················  ····················  ····················
I   2814 hCoV-19/Canada/ON-PHL-20-01222/2020|EPI_ISL_548844   ····················  ······A·············  ····················  ····················  ····················  ····················
I   2815 hCoV-19/Canada/ON-PHL-20-01223/2020|EPI_ISL_548845   ····················  ····················  ····················  ····················  ····················  ····················
I   2816 hCoV-19/Canada/ON-PHL-20-01224/2020|EPI_ISL_548846   ····················  ····················  ····················  ····················  NNN·················  ····················   1
I   2824 hCoV-19/USA/TX-HMH-4758/2020|EPI_ISL_544309|2020-0   ····················  ····················  ····················  ····················  ····················  ····················
I   2826 hCoV-19/USA/TX-HMH-476/2020|EPI_ISL_544311|2020-04   ····················  ····················  ····················  ····················  ····················  ····················
I   2828 hCoV-19/USA/TX-HMH-4764/2020|EPI_ISL_544314|2020-0   ····················  ····················  ····················  ····················  ····················  ····················
I   2829 hCoV-19/USA/TX-HMH-4844/2020|EPI_ISL_544370|2020-0   ····················  ·············A······  ····················  ····················  ····················  ····················
I   2830 hCoV-19/USA/TX-HMH-4845/2020|EPI_ISL_544371|2020-0   ····················  ····················  ····················  ····················  ····················  ····················
I   2831 hCoV-19/USA/TX-HMH-4846/2020|EPI_ISL_544372|2020-0   ····················  ····················  ····················  ····················  ····················  ····················
I   2832 hCoV-19/USA/TX-HMH-4847/2020|EPI_ISL_544373|2020-0   ····················  ····················  ····················  ····················  ····················  ····················
I   2833 hCoV-19/USA/TX-HMH-4848/2020|EPI_ISL_544374|2020-0   ····················  ····················  ····················  ····················  ····················  ····················
I   2834 hCoV-19/USA/TX-HMH-4852/2020|EPI_ISL_544376|2020-0   ····················  ····················  ····················  ····················  ····················  ····················
I   2835 hCoV-19/USA/TX-HMH-4854/2020|EPI_ISL_544377|2020-0   ····················  ····················  ····················  ····················  ····················  ····················
I   2836 hCoV-19/USA/TX-HMH-4858/2020|EPI_ISL_544380|2020-0   ····················  ····················  ····················  ····················  ····················  ·······NN···········
```

| | | | | | | | | |
|---|---|---|---|---|---|---|---|---|
| I | 2837 | hCoV-19/USA/TX-HMH-486/2020\|EPI_ISL_544381\|2020-04 | ···················· | ···················· | ···················· | ···················· | ···················· | ···················· |
| I | 2838 | hCoV-19/USA/TX-HMH-4862/2020\|EPI_ISL_544384\|2020-0 | ···················· | ·············A······ | ···················· | ···················· | ···················· | ···················· |
| I | 2839 | hCoV-19/USA/TX-HMH-4864/2020\|EPI_ISL_544386\|2020-0 | ···················· | ···················· | ···················· | ···················· | ···················· | ···················· |
| I | 2840 | hCoV-19/USA/TX-HMH-4865/2020\|EPI_ISL_544387\|2020-0 | ···················· | ···················· | ···················· | ···················· | ···················· | ···················· |
| I | 2841 | hCoV-19/USA/TX-HMH-4867/2020\|EPI_ISL_544389\|2020-0 | ···················· | ···················· | ···················· | ···················· | ···················· | ···················· |
| I | 2842 | hCoV-19/USA/TX-HMH-4868/2020\|EPI_ISL_544390\|2020-0 | ···················· | ···················· | ···················· | ···················· | ···················· | ···················· |
| I | 2843 | hCoV-19/USA/TX-HMH-4869/2020\|EPI_ISL_544391\|2020-0 | ···················· | ···················· | ···················· | ···················· | ···················· | ···················· |
| I | 2844 | hCoV-19/USA/TX-HMH-487/2020\|EPI_ISL_544392\|2020-04 | ···················· | ···················· | ···················· | ···················· | ···················· | ···················· |
| I | 2846 | hCoV-19/USA/TX-HMH-4875/2020\|EPI_ISL_544394\|2020-0 | ···················· | ···················· | ···················· | ···················· | ···················· | ···················· |
| I | 2847 | hCoV-19/USA/TX-HMH-4878/2020\|EPI_ISL_544396\|2020-0 | ···················· | ···················· | ···················· | ···················· | ···················· | ···A················ |
| I | 2848 | hCoV-19/USA/TX-HMH-4879/2020\|EPI_ISL_544397\|2020-0 | ···················· | ···················· | ···················· | ···················· | ···················· | ···················· |
| I | 2849 | hCoV-19/USA/TX-HMH-488/2020\|EPI_ISL_544398\|2020-04 | ···················· | ···················· | ···················· | ···················· | ···················· | ···················· |
| I | 2851 | hCoV-19/USA/TX-HMH-7747/2020\|EPI_ISL_545614\|2020-0 | ···················· | ···················· | ···················· | ···················· | ···················· | ···················· |
| I | 2852 | hCoV-19/USA/TX-HMH-7751/2020\|EPI_ISL_545615\|2020-0 | ···················· | ···················· | ···················· | ···················· | ···················· | ···················· |
| I | 2853 | hCoV-19/USA/TX-HMH-7757/2020\|EPI_ISL_545619\|2020-0 | ···················· | ···················· | ···················· | ···················· | ···················· | ···················· |
| I | 2854 | hCoV-19/USA/TX-HMH-7759/2020\|EPI_ISL_545621\|2020-0 | ···················· | ·············A······ | ···················· | ···················· | ···················· | ···················· |
| I | 2856 | hCoV-19/USA/TX-HMH-7768/2020\|EPI_ISL_545625\|2020-0 | ···················· | ···················· | ···················· | ···················· | ···················· | ···················· |
| I | 2857 | hCoV-19/USA/TX-HMH-7769/2020\|EPI_ISL_545626\|2020-0 | ···················· | ···················· | ···················· | ···················· | ···················· | ···················· |
| I | 2858 | hCoV-19/USA/TX-HMH-777/2020\|EPI_ISL_545627\|2020-04 | ···················· | ···················· | ···················· | ···················· | ···················· | ···················· |
| I | 2859 | hCoV-19/USA/TX-HMH-7774/2020\|EPI_ISL_545630\|2020-0 | ···················· | ···················· | ···················· | ···················· | ···················· | ···················· |
| I | 2860 | hCoV-19/USA/TX-HMH-7780/2020\|EPI_ISL_545633\|2020-0 | ···················· | ···················· | ···················· | ···················· | ···················· | ···················· |
| I | 2861 | hCoV-19/USA/TX-HMH-7782/2020\|EPI_ISL_545634\|2020-0 | ···················· | ···················· | ···················· | ···················· | ···················· | ···················· |
| I | 2862 | hCoV-19/USA/TX-HMH-7787/2020\|EPI_ISL_545636\|2020-0 | ···················· | ···················· | ···················· | ···················· | ···················· | ···················· |
| I | 2863 | hCoV-19/USA/TX-HMH-779/2020\|EPI_ISL_545637\|2020-04 | ···················· | ···················· | ···················· | ···················· | ···················· | ···················· |
| I | 2864 | hCoV-19/USA/TX-HMH-7798/2020\|EPI_ISL_545638\|2020-0 | ···················· | ···················· | ···················· | ···················· | ···················· | ···················· |
| I | 2865 | hCoV-19/USA/TX-HMH-7799/2020\|EPI_ISL_545639\|2020-0 | ···················· | ·A·················· | ···················· | ···················· | ···················· | ···················· |
| I | 2866 | hCoV-19/USA/TX-HMH-780/2020\|EPI_ISL_545640\|2020-04 | ···················· | ···················· | ···················· | ···················· | ···················· | ···················· |
| I | 2867 | hCoV-19/USA/TX-HMH-7808/2020\|EPI_ISL_545644\|2020-0 | ···················· | ···················· | ···················· | ···················· | ···················· | ···················· |
| I | 2868 | hCoV-19/USA/TX-HMH-781/2020\|EPI_ISL_545645\|2020-04 | ···················· | ···················· | ···················· | ···················· | ···················· | ···················· |
| I | 2869 | hCoV-19/USA/TX-HMH-6580/2020\|EPI_ISL_546100\|2020-0 | ···················· | ·············A······ | ···················· | ···················· | ···················· | ···················· |
| I | 2870 | hCoV-19/USA/TX-HMH-6581/2020\|EPI_ISL_546101\|2020-0 | ···················· | ···················· | ···················· | ···················· | ···················· | ·······N············ |
| I | 2871 | hCoV-19/USA/TX-HMH-6583/2020\|EPI_ISL_546102\|2020-0 | ···················· | ···················· | ···················· | ···················· | ···················· | ···················· |
| I | 2872 | hCoV-19/USA/TX-HMH-6585/2020\|EPI_ISL_546103\|2020-0 | ···················· | ···················· | ···················· | ···················· | ···················· | ···················· |
| I | 2873 | hCoV-19/USA/TX-HMH-6587/2020\|EPI_ISL_546105\|2020-0 | ···················· | ···················· | ···················· | ···················· | ···················· | ···················· |
| I | 2874 | hCoV-19/USA/TX-HMH-6588/2020\|EPI_ISL_546106\|2020-0 | ···················· | ·············A······ | ···················· | ···················· | ···················· | ···················· |
| I | 2875 | hCoV-19/USA/TX-HMH-6590/2020\|EPI_ISL_546108\|2020-0 | ···················· | ···················· | ···················· | ···················· | ···················· | ···················· |
| I | 2876 | hCoV-19/USA/TX-HMH-6591/2020\|EPI_ISL_546109\|2020-0 | ···················· | ···················· | ···················· | ···················· | ···················· | ···················· |
| I | 2877 | hCoV-19/USA/TX-HMH-6592/2020\|EPI_ISL_546110\|2020-0 | ···················· | ···················· | ···················· | ···················· | ···················· | ···················· |
| I | 2878 | hCoV-19/USA/TX-HMH-6593/2020\|EPI_ISL_546111\|2020-0 | ···················· | ·············A······ | ···················· | ···················· | ···················· | ···················· |
| I | 2879 | hCoV-19/USA/TX-HMH-6595/2020\|EPI_ISL_546113\|2020-0 | ···················· | ···················· | ···················· | ···················· | ···················· | ···················· |
| I | 2880 | hCoV-19/USA/TX-HMH-6596/2020\|EPI_ISL_546114\|2020-0 | ···················· | ···················· | ···················· | ···················· | ···················· | ···················· |
| I | 2881 | hCoV-19/USA/TX-HMH-6598/2020\|EPI_ISL_546116\|2020-0 | ···················· | ···················· | ···················· | ···················· | ···················· | ···················· |
| I | 2882 | hCoV-19/USA/TX-HMH-6599/2020\|EPI_ISL_546117\|2020-0 | ···················· | ·A·················· | ···················· | ···················· | ···················· | ···················· |
| I | 2883 | hCoV-19/USA/TX-HMH-6602/2020\|EPI_ISL_546119\|2020-0 | ···················· | ···················· | ···················· | ···················· | ···················· | ···················· |
| I | 2884 | hCoV-19/USA/TX-HMH-6615/2020\|EPI_ISL_546120\|2020-0 | ···················· | ···················· | ···················· | ···················· | ···················· | ···················· |
| I | 2885 | hCoV-19/USA/TX-HMH-6616/2020\|EPI_ISL_546121\|2020-0 | ···················· | ···················· | ···················· | ···················· | ···················· | ···················· |
| I | 2886 | hCoV-19/USA/TX-HMH-6620/2020\|EPI_ISL_546123\|2020-0 | ···················· | ···················· | ···A················ | ···················· | ···················· | ···················· |
| I | 2887 | hCoV-19/USA/TX-HMH-6625/2020\|EPI_ISL_546127\|2020-0 | ···················· | ···················· | ···················· | ···················· | ···················· | ······NN············ |
| I | 2888 | hCoV-19/USA/TX-HMH-6626/2020\|EPI_ISL_546128\|2020-0 | ···················· | ·············A······ | ···················· | ···················· | ···················· | ···················· |
| I | 2889 | hCoV-19/USA/TX-HMH-6627/2020\|EPI_ISL_546129\|2020-0 | ···················· | ···················· | ···················· | ···················· | ···················· | ···················· |
| I | 2891 | hCoV-19/USA/TX-HMH-6638/2020\|EPI_ISL_546136\|2020-0 | ···················· | ···················· | ···················· | ···················· | ···················· | ···················· |
| I | 2892 | hCoV-19/USA/TX-HMH-6639/2020\|EPI_ISL_546137\|2020-0 | ···················· | ···················· | ···················· | ···················· | ···················· | ···················· |
| I | 2893 | hCoV-19/USA/TX-HMH-6640/2020\|EPI_ISL_546138\|2020-0 | ···················· | ·············A······ | ···················· | ···················· | ···················· | ···················· |
| I | 2894 | hCoV-19/USA/TX-HMH-6642/2020\|EPI_ISL_546139\|2020-0 | ···················· | ·············A······ | ···················· | ···················· | ···················· | ···················· |
| I | 2899 | hCoV-19/USA/TX-HMH-6651/2020\|EPI_ISL_546146\|2020-0 | ···················· | ···················· | ···················· | ···················· | ···················· | ···················· |
| I | 2900 | hCoV-19/USA/TX-HMH-6652/2020\|EPI_ISL_546147\|2020-0 | ···················· | ···················· | ···················· | ···················· | ···················· | ···················· |
| I | 2901 | hCoV-19/USA/TX-HMH-6653/2020\|EPI_ISL_546148\|2020-0 | ···················· | ···················· | ···················· | ···················· | ···················· | ···················· |
| I | 2906 | hCoV-19/USA/TX-HMH-5695/2020\|EPI_ISL_546949\|2020-0 | ···················· | ···················· | ···················· | ···················· | ···················· | ···················· |
| I | 2907 | hCoV-19/USA/TX-HMH-5697/2020\|EPI_ISL_546950\|2020-0 | ···················· | ···················· | ···················· | ···················· | ···················· | ···················· |
| I | 2909 | hCoV-19/USA/TX-HMH-570/2020\|EPI_ISL_546952\|2020-04 | ···················· | ···················· | ···················· | ···················· | ···················· | ···················· |
| I | 2912 | hCoV-19/USA/TX-HMH-5703/2020\|EPI_ISL_546955\|2020-0 | ···················· | ·············A······ | ···················· | ···················· | ···················· | ···················· |
| I | 2913 | hCoV-19/USA/TX-HMH-5706/2020\|EPI_ISL_546956\|2020-0 | ···················· | ·············A······ | ···················· | ···················· | ···················· | ···················· |
| I | 2914 | hCoV-19/USA/TX-HMH-5707/2020\|EPI_ISL_546957\|2020-0 | ···················· | ·············A······ | ···················· | ···················· | ···················· | ···················· |
| I | 2921 | hCoV-19/USA/TX-HMH-5721/2020\|EPI_ISL_546967\|2020-0 | ···················· | ···················· | ···················· | ···················· | ···················· | ···················· |
| I | 2922 | hCoV-19/USA/TX-HMH-5722/2020\|EPI_ISL_546968\|2020-0 | ···················· | ···················· | ···················· | ···················· | ···················· | ···················· |
| I | 2923 | hCoV-19/USA/TX-HMH-5731/2020\|EPI_ISL_546973\|2020-0 | ···················· | ···················· | ···················· | ···················· | ···················· | ···················· |
| I | 2924 | hCoV-19/USA/TX-HMH-5732/2020\|EPI_ISL_546974\|2020-0 | ···················· | ···················· | ···················· | ···················· | ···················· | ···················· |
| I | 2927 | hCoV-19/USA/TX-HMH-6340/2020\|EPI_ISL_547402\|2020-0 | ···················· | ···················· | ···················· | ···················· | ···················· | ···················· |
| I | 2928 | hCoV-19/USA/TX-HMH-6341/2020\|EPI_ISL_547403\|2020-0 | ···················· | ···················· | ···················· | ···················· | ···················· | N··················· |
| I | 2929 | hCoV-19/USA/TX-HMH-6343/2020\|EPI_ISL_547404\|2020-0 | ···················· | ···················· | ···················· | ···················· | ··············N····· | ···················· |
| I | 2930 | hCoV-19/USA/TX-HMH-6346/2020\|EPI_ISL_547405\|2020-0 | ···················· | ···················· | ···················· | ···················· | ···················· | ···················· |
| I | 2931 | hCoV-19/USA/TX-HMH-6350/2020\|EPI_ISL_547408\|2020-0 | ···················· | ·············A······ | ···················· | ···················· | ···················· | ···················· |
| I | 2932 | hCoV-19/USA/TX-HMH-6351/2020\|EPI_ISL_547409\|2020-0 | ···················· | ···················· | ···················· | ···················· | ···················· | ···················· |
| I | 2933 | hCoV-19/USA/TX-HMH-6352/2020\|EPI_ISL_547410\|2020-0 | ···················· | ·············A······ | ···················· | ···················· | ···················· | ···················· |
| I | 2934 | hCoV-19/USA/TX-HMH-6354/2020\|EPI_ISL_547411\|2020-0 | ···················· | ···················· | ···················· | ···················· | ···················· | ···················· |
| I | 2936 | hCoV-19/USA/TX-HMH-6356/2020\|EPI_ISL_547413\|2020-0 | ···················· | ·············A······ | ···················· | ···················· | ···················· | ···················· |
| I | 2938 | hCoV-19/USA/TX-HMH-636/2020\|EPI_ISL_547416\|2020-04 | ···················· | ···················· | ···················· | ···················· | ···················· | ···················· |
| I | 2941 | hCoV-19/USA/TX-HMH-6365/2020\|EPI_ISL_547420\|2020-0 | ···················· | ···················· | ···················· | ···················· | ···················· | ···················· |
| I | 2942 | hCoV-19/USA/TX-HMH-6366/2020\|EPI_ISL_547421\|2020-0 | ···················· | ···················· | ···················· | ···················· | ···················· | ···················· |
| I | 2943 | hCoV-19/USA/TX-HMH-6368/2020\|EPI_ISL_547423\|2020-0 | ···················· | ···················· | ···················· | ···················· | ···················· | NN·N················ |

1

| | | | | | | | | |
|---|---|---|---|---|---|---|---|---|
| I | 2944 | hCoV-19/USA/TX-HMH-6370/2020\|EPI_ISL_547424\|2020-0 | .................... | .................... | .................... | .................... | .................... | .................... |
| I | 2945 | hCoV-19/USA/TX-HMH-6372/2020\|EPI_ISL_547426\|2020-0 | .................... | .................... | .................... | .................... | .................... | .................... |
| I | 2946 | hCoV-19/USA/TX-HMH-6373/2020\|EPI_ISL_547427\|2020-0 | .................... | .................... | .................... | .................... | .................... | .................... |
| I | 2947 | hCoV-19/USA/TX-HMH-6381/2020\|EPI_ISL_547431\|2020-0 | .................... | .................... | .................... | .................... | .................... | .................... |
| I | 2948 | hCoV-19/USA/TX-HMH-6382/2020\|EPI_ISL_547432\|2020-0 | .................... | .................... | .................... | .................... | .................... | .................... |
| I | 2949 | hCoV-19/Gibraltar/203160593/2020\|EPI_ISL_547433\|20 | .................... | .................... | .................... | .................... | .................... | .................... |
| I | 2950 | hCoV-19/Gibraltar/203160594/2020\|EPI_ISL_547434\|20 | .................... | .................... | .................... | .................... | .................... | .................... |
| I | 2951 | hCoV-19/Gibraltar/203160595/2020\|EPI_ISL_547435\|20 | .................... | .................... | .................... | .................... | .................... | .................... |
| I | 2952 | hCoV-19/Gibraltar/203260482/2020\|EPI_ISL_547436\|20 | .................... | .................... | .................... | .................... | .................... | .................... |
| I | 2953 | hCoV-19/Gibraltar/203360721/2020\|EPI_ISL_547437\|20 | .................... | .................... | .................... | .................... | .................... | .................... |
| I | 2954 | hCoV-19/Gibraltar/203360722/2020\|EPI_ISL_547438\|20 | .................... | .................... | .................... | .................... | .................... | .................... |
| I | 2955 | hCoV-19/Gibraltar/203360724/2020\|EPI_ISL_547439\|20 | .................... | .................... | .................... | .................... | .................... | .................... |
| I | 2956 | hCoV-19/Gibraltar/203360725/2020\|EPI_ISL_547440\|20 | .................... | .................... | .................... | .................... | .................... | .................... |
| I | 2957 | hCoV-19/Gibraltar/203360726/2020\|EPI_ISL_547441\|20 | .................... | .A.................. | .................... | .................... | .................... | .................... |
| I | 2958 | hCoV-19/Gibraltar/203360727/2020\|EPI_ISL_547442\|20 | .................... | .A.................. | .................... | .................... | .................... | .................... |
| I | 2959 | hCoV-19/Gibraltar/203360728/2020\|EPI_ISL_547443\|20 | .................... | .................... | .................... | .................... | .................... | .................... |
| I | 2960 | hCoV-19/Gibraltar/203360729/2020\|EPI_ISL_547444\|20 | .................... | .................... | .................... | .................... | .................... | .................... |
| I | 2961 | hCoV-19/Aruba/AW-RIVM-10101/2020\|EPI_ISL_547445\|20 | .................... | .................... | .................... | .................... | .................... | .................... |
| I | 2962 | hCoV-19/Aruba/AW-RIVM-10113/2020\|EPI_ISL_547446\|20 | .................... | .................... | .................... | .................... | .................... | .................... |
| I | 2963 | hCoV-19/Aruba/AW-RIVM-10114/2020\|EPI_ISL_547447\|20 | .................... | .................... | .................... | .................... | .................... | .................... |
| I | 2964 | hCoV-19/Aruba/AW-RIVM-10115/2020\|EPI_ISL_547448\|20 | .................... | .................... | .................... | .................... | .................... | .................... |
| I | 2965 | hCoV-19/Aruba/AW-RIVM-10122/2020\|EPI_ISL_547449\|20 | .................... | .................... | .................... | .................... | .................... | .................... |
| I | 2966 | hCoV-19/Curacao/CW-RIVM-10092/2020\|EPI_ISL_547450\| | .................... | .................... | .................... | .................... | .................... | .................... |
| I | 2967 | hCoV-19/Curacao/CW-RIVM-10093/2020\|EPI_ISL_547451\| | .................... | .................... | .................... | .................... | .................... | .................... |
| I | 2968 | hCoV-19/Curacao/CW-RIVM-10094/2020\|EPI_ISL_547452\| | .................... | .................... | .................... | .................... | .................... | .................... |
| I | 2969 | hCoV-19/Curacao/CW-RIVM-10100/2020\|EPI_ISL_547453\| | .................... | .................... | .................... | .................... | .................... | .................... |
| I | 2970 | hCoV-19/Netherlands/FL-RIVM-10132/2020\|EPI_ISL_547 | .................... | .................... | .................... | .................... | .................... | .................... |
| I | 2971 | hCoV-19/Netherlands/FL-RIVM-10142/2020\|EPI_ISL_547 | .................... | .................... | .................... | .................... | .................... | .................... |
| I | 2972 | hCoV-19/Netherlands/FL-RIVM-10143/2020\|EPI_ISL_547 | .................... | .................... | .................... | .................... | .................... | .................... |
| I | 2973 | hCoV-19/Netherlands/FR-RIVM-10083/2020\|EPI_ISL_547 | .................... | .................... | .................... | .................... | .................... | .................... |
| I | 2974 | hCoV-19/Netherlands/FR-RIVM-10084/2020\|EPI_ISL_547 | .................... | .................... | .................... | .................... | .................... | .................... |
| I | 2975 | hCoV-19/Netherlands/GE-RIVM-10075/2020\|EPI_ISL_547 | .................... | .................... | .................... | .................... | .................... | .................... |
| I | 2976 | hCoV-19/Netherlands/GE-RIVM-10088/2020\|EPI_ISL_547 | .................... | .................... | .................... | .................... | .................... | .................... |
| I | 2977 | hCoV-19/Netherlands/GE-RIVM-10095/2020\|EPI_ISL_547 | .................... | .................... | .................... | .................... | .................... | .................... |
| I | 2978 | hCoV-19/Netherlands/GE-RIVM-10098/2020\|EPI_ISL_547 | .................... | .................... | .................... | .................... | .................... | .................... |
| I | 2979 | hCoV-19/Netherlands/GE-RIVM-10111/2020\|EPI_ISL_547 | .................... | .................... | .................... | .................... | .................... | .................... |
| I | 2980 | hCoV-19/Netherlands/GE-RIVM-10112/2020\|EPI_ISL_547 | .................... | .................... | .................... | .................... | .................... | .................... |
| I | 2981 | hCoV-19/Netherlands/GE-RIVM-10124/2020\|EPI_ISL_547 | .................... | .................... | .................... | .................... | .................... | .................... |
| I | 2982 | hCoV-19/Netherlands/GR-RIVM-10074/2020\|EPI_ISL_547 | .................... | .................... | .................... | .................... | .................... | .................... |
| I | 2983 | hCoV-19/Netherlands/GR-RIVM-10081/2020\|EPI_ISL_547 | .................... | .................... | .................... | .................... | .................... | .................... |
| I | 2984 | hCoV-19/Netherlands/GR-RIVM-10089/2020\|EPI_ISL_547 | .................... | .................... | .................... | .................... | .................... | .................... |
| I | 2985 | hCoV-19/Netherlands/LI-RIVM-10096/2020\|EPI_ISL_547 | .................... | .................... | .................... | .................... | .................... | .................... |
| I | 2986 | hCoV-19/Netherlands/LI-RIVM-10097/2020\|EPI_ISL_547 | .................... | .................... | .................... | .................... | .................... | .................... |
| I | 2987 | hCoV-19/Netherlands/LI-RIVM-10105/2020\|EPI_ISL_547 | .................... | .................... | .................... | .................... | .................... | .................... |
| I | 2988 | hCoV-19/Netherlands/LI-RIVM-10106/2020\|EPI_ISL_547 | .................... | .................... | .................... | .................... | .................... | .................... |
| I | 2989 | hCoV-19/Netherlands/LI-RIVM-10119/2020\|EPI_ISL_547 | .................... | .................... | .................... | .................... | .................... | .................... |
| I | 2990 | hCoV-19/Netherlands/LI-RIVM-10130/2020\|EPI_ISL_547 | .................... | .................... | .................... | .................... | .................... | .................... |
| I | 2991 | hCoV-19/Netherlands/LI-RIVM-10131/2020\|EPI_ISL_547 | .................... | .................... | .................... | .................... | .................... | .................... |
| I | 2992 | hCoV-19/Netherlands/LI-RIVM-10146/2020\|EPI_ISL_547 | .................... | .................... | .................... | .................... | .................... | .................... |
| I | 2993 | hCoV-19/Netherlands/LI-RIVM-10147/2020\|EPI_ISL_547 | .................... | .................... | .................... | .................... | .................... | .................... |
| I | 2994 | hCoV-19/Netherlands/LI-RIVM-10150/2020\|EPI_ISL_547 | .................... | .................... | .................... | .................... | .................... | .................... |
| I | 2995 | hCoV-19/Netherlands/LI-RIVM-10151/2020\|EPI_ISL_547 | .................... | .................... | .................... | .................... | .................... | .................... |
| I | 2996 | hCoV-19/New Zealand/20CV0143/2020\|EPI_ISL_548021\|2 | .................... | .................... | .................... | .................... | .................... | .................... |
| I | 2997 | hCoV-19/New Zealand/20CV0144/2020\|EPI_ISL_548022\|2 | .................... | .................... | .................... | .................... | .................... | .................... |
| I | 2998 | hCoV-19/New Zealand/20CV0149/2020\|EPI_ISL_548025\|2 | .................... | .................... | .................... | .................... | .................... | .................... |
| I | 2999 | hCoV-19/New Zealand/20CV0156/2020\|EPI_ISL_548030\|2 | .................... | .................... | .................... | .................... | .................... | .................... |
| I | 3000 | hCoV-19/New Zealand/20CV0169/2020\|EPI_ISL_548040\|2 | .................... | .................... | .................... | .................... | .................... | .................... |
| I | 3001 | hCoV-19/New Zealand/20CV0170/2020\|EPI_ISL_548041\|2 | .................... | .................... | .................... | .................... | .................... | .................... |
| I | 3002 | hCoV-19/New Zealand/20CV0176/2020\|EPI_ISL_548045\|2 | .................... | .................... | .................... | .................... | .................... | .................... |
| I | 3003 | hCoV-19/New Zealand/20CV0177/2020\|EPI_ISL_548046\|2 | .................... | .................... | .................... | .................... | .................... | .................... |
| I | 3004 | hCoV-19/New Zealand/20CV0178/2020\|EPI_ISL_548047\|2 | .................... | .................... | .................... | .................... | .................... | .................... |
| I | 3005 | hCoV-19/New Zealand/20CV0180/2020\|EPI_ISL_548048\|2 | .................... | .................... | .................... | .................... | .................... | .................... |
| I | 3006 | hCoV-19/New Zealand/20CV0181/2020\|EPI_ISL_548049\|2 | .................... | .................... | .................... | .................... | .................... | .................... |
| I | 3007 | hCoV-19/Canada/ON-PHL-20-00049/2020\|EPI_ISL_548726 | .................... | .................... | .................... | .................... | .................... | .................... |
| I | 3008 | hCoV-19/Canada/ON-PHL-20-00050/2020\|EPI_ISL_548727 | .................... | .................... | .................... | .................... | .................... | .................... |
| I | 3009 | hCoV-19/Canada/ON-PHL-20-00051/2020\|EPI_ISL_548728 | .................... | .................... | .................... | .................... | .................... | .................... |
| I | 3010 | hCoV-19/Canada/ON-PHL-20-00052/2020\|EPI_ISL_548729 | .................... | .................... | .................... | .................... | .................... | .................... |
| I | 3011 | hCoV-19/Canada/ON-PHL-20-00053/2020\|EPI_ISL_548730 | .................... | .................... | .................... | .................... | .................... | .................... |
| I | 3012 | hCoV-19/Canada/ON-PHL-20-00054/2020\|EPI_ISL_548731 | .................... | .................... | .................... | .................... | .................... | .................... |
| I | 3013 | hCoV-19/Canada/ON-PHL-20-00055/2020\|EPI_ISL_548732 | .................... | .................... | .................... | .................... | .................... | .................... |
| I | 3014 | hCoV-19/Canada/ON-PHL-20-00056/2020\|EPI_ISL_548733 | .................... | .................... | .................... | .................... | .................... | .................... |
| I | 3015 | hCoV-19/Canada/ON-PHL-20-00057/2020\|EPI_ISL_548734 | .................... | .................... | .................... | .................... | .................... | .................... |
| I | 3016 | hCoV-19/Canada/ON-PHL-20-00059/2020\|EPI_ISL_548735 | .................... | .................... | .................... | .................... | .................... | .................... |
| I | 3017 | hCoV-19/Canada/ON-PHL-20-00061/2020\|EPI_ISL_548737 | .................... | .................... | .................... | .................... | .................... | .................... |
| I | 3018 | hCoV-19/Canada/ON-PHL-20-00062/2020\|EPI_ISL_548738 | .................... | .................... | .................... | .................... | .................... | .................... |
| I | 3019 | hCoV-19/Canada/ON-PHL-20-00064/2020\|EPI_ISL_548740 | .................... | .................... | .................... | .................... | .................... | .................... |
| I | 3020 | hCoV-19/Canada/ON-PHL-20-00065/2020\|EPI_ISL_548741 | .................... | .................... | .................... | .................... | .................... | .................... |
| I | 3021 | hCoV-19/Canada/ON-PHL-20-00066/2020\|EPI_ISL_548742 | .................... | .................... | .................... | .................... | .................... | .................... |
| I | 3022 | hCoV-19/Canada/ON-PHL-20-00068/2020\|EPI_ISL_548743 | .................... | .................... | .................... | .................... | .................... | .................... |
| I | 3023 | hCoV-19/Canada/ON-PHL-20-00069/2020\|EPI_ISL_548744 | .................... | .................... | .................... | .................... | .................... | .................... |

| | | | | | | | | |
|---|---|---|---|---|---|---|---|---|
| I | 3024 | hCoV-19/Canada/ON-PHL-20-00070/2020\|EPI_ISL_548745 | .................... | .................... | .................... | .................... | .................... | .................... |
| I | 3025 | hCoV-19/Canada/ON-PHL-20-00071/2020\|EPI_ISL_548746 | .................... | .................... | .................... | .................... | .................... | .................... |
| I | 3026 | hCoV-19/Canada/ON-PHL-20-00072/2020\|EPI_ISL_548747 | .................... | .................... | .................... | .................... | .................... | .................... |
| I | 3027 | hCoV-19/Canada/ON-PHL-20-00075/2020\|EPI_ISL_548750 | .................... | .................... | .................... | .................... | .................... | .................... |
| I | 3028 | hCoV-19/Canada/ON-PHL-20-00076/2020\|EPI_ISL_548751 | .................... | .................... | .................... | .................... | .................... | .................... |
| I | 3029 | hCoV-19/Canada/ON-PHL-20-00077/2020\|EPI_ISL_548752 | .................... | .................... | .................... | .................... | .................... | .................... |
| I | 3030 | hCoV-19/Canada/ON-PHL-20-00078/2020\|EPI_ISL_548753 | .................... | .................... | .................... | .................... | .................... | .................... |
| I | 3031 | hCoV-19/Canada/ON-PHL-20-00079/2020\|EPI_ISL_548754 | .................... | .................... | .................... | .................... | .................... | .................... |
| I | 3032 | hCoV-19/Canada/ON-PHL-20-00080/2020\|EPI_ISL_548755 | .................... | .................... | .................... | .................... | .................... | .................... |
| I | 3033 | hCoV-19/Canada/ON-PHL-20-00086/2020\|EPI_ISL_548759 | .................... | .................... | .................... | .................... | .................... | .................... |
| I | 3034 | hCoV-19/Canada/ON-PHL-20-00087/2020\|EPI_ISL_548760 | .................... | .................... | .................... | .................... | .................... | .................... |
| I | 3035 | hCoV-19/Canada/ON-PHL-20-00088/2020\|EPI_ISL_548761 | .................... | .................... | .................... | .................... | .................... | .................... |
| I | 3036 | hCoV-19/Canada/ON-PHL-20-00169/2020\|EPI_ISL_548762 | .................... | .................... | .................... | .................... | .................... | .................... |
| I | 3037 | hCoV-19/Canada/ON-PHL-20-00170/2020\|EPI_ISL_548763 | .................... | .................... | .................... | .................... | .................... | .................... |
| I | 3038 | hCoV-19/Canada/ON-PHL-20-00171/2020\|EPI_ISL_548764 | .................... | .................... | .................... | .................... | .................... | .................... |
| I | 3039 | hCoV-19/Canada/ON-PHL-20-00172/2020\|EPI_ISL_548765 | .................... | .................... | .................... | .................... | .................... | .................... |
| I | 3040 | hCoV-19/Canada/ON-PHL-20-00173/2020\|EPI_ISL_548766 | .................... | .................... | .................... | .................... | .................... | .................... |
| I | 3041 | hCoV-19/Canada/ON-PHL-20-00174/2020\|EPI_ISL_548767 | .................... | .................... | .................... | .................... | .................... | .................... |
| I | 3042 | hCoV-19/Canada/ON-PHL-20-00233/2020\|EPI_ISL_548768 | .................... | .................... | .................... | .................... | .................... | .................... |
| I | 3043 | hCoV-19/Canada/ON-PHL-20-00234/2020\|EPI_ISL_548769 | .................... | .................... | .................... | .................... | .................... | .................... |
| I | 3044 | hCoV-19/Canada/ON-PHL-20-00235/2020\|EPI_ISL_548770 | .................... | .................... | .................... | .................... | .................... | .................... |
| I | 3045 | hCoV-19/Canada/ON-PHL-20-00236/2020\|EPI_ISL_548771 | .................... | .................... | .................... | .................... | .................... | .................... |
| I | 3046 | hCoV-19/Canada/ON-PHL-20-00237/2020\|EPI_ISL_548772 | .................... | .................... | .................... | .................... | .................... | .................... |
| I | 3047 | hCoV-19/Canada/ON-PHL-20-00238/2020\|EPI_ISL_548773 | .................... | .................... | .................... | .................... | .................... | .................... |
| I | 3048 | hCoV-19/Canada/ON-PHL-20-00320/2020\|EPI_ISL_548774 | .................... | .................... | .................... | .................... | .................... | .................... |
| I | 3049 | hCoV-19/Canada/ON-PHL-20-00384/2020\|EPI_ISL_548775 | .................... | .................... | .................... | .................... | .................... | .................... |
| I | 3050 | hCoV-19/Canada/ON-PHL-20-00385/2020\|EPI_ISL_548776 | .................... | .................... | .................... | .................... | .................... | .................... |
| I | 3051 | hCoV-19/Canada/ON-PHL-20-00386/2020\|EPI_ISL_548777 | .................... | .................... | .................... | .................... | .................... | .................... |
| I | 3052 | hCoV-19/Canada/ON-PHL-20-00397/2020\|EPI_ISL_548778 | .................... | .................... | .................... | .................... | .................... | .................... |
| I | 3053 | hCoV-19/Canada/ON-PHL-20-00820/2020\|EPI_ISL_548779 | .................... | .................... | .................... | .................... | .................... | .................... |
| I | 3054 | hCoV-19/Canada/ON-PHL-20-00987/2020\|EPI_ISL_548780 | .................... | .................... | .................... | .................... | .................... | .................... |
| I | 3055 | hCoV-19/Canada/ON-PHL-20-01158/2020\|EPI_ISL_548781 | .................... | .................... | .................... | .................... | .................... | .................... |
| I | 3056 | hCoV-19/Canada/ON-PHL-20-01160/2020\|EPI_ISL_548782 | .................... | .................... | .................... | .................... | .................... | .................... |
| I | 3057 | hCoV-19/Canada/ON-PHL-20-01161/2020\|EPI_ISL_548783 | .................... | .................... | .................... | .................... | .................... | .................... |
| I | 3058 | hCoV-19/Canada/ON-PHL-20-01162/2020\|EPI_ISL_548784 | .................... | .................... | .................... | .................... | .................... | .................... |
| I | 3060 | hCoV-19/Canada/ON-PHL-20-01164/2020\|EPI_ISL_548786 | .................... | .................... | .................... | .................... | .................... | .................... |
| I | 3061 | hCoV-19/Canada/ON-PHL-20-01165/2020\|EPI_ISL_548787 | .................... | .................... | .................... | .................... | .................... | .................... |
| I | 3062 | hCoV-19/Canada/ON-PHL-20-01166/2020\|EPI_ISL_548788 | .................... | .................... | .................... | .................... | .................... | .................... |
| I | 3063 | hCoV-19/Canada/ON-PHL-20-01167/2020\|EPI_ISL_548789 | .................... | .................... | .................... | .................... | .................... | .................... |
| I | 3064 | hCoV-19/Canada/ON-PHL-20-01168/2020\|EPI_ISL_548790 | .................... | .................... | .................... | .................... | .................... | .................... |
| I | 3065 | hCoV-19/Canada/ON-PHL-20-01169/2020\|EPI_ISL_548791 | .................... | .................... | .................... | .................... | .................... | .................... |
| I | 3066 | hCoV-19/Canada/ON-PHL-20-01170/2020\|EPI_ISL_548792 | .................... | .................... | .................... | .................... | .................... | .................... |
| I | 3067 | hCoV-19/Canada/ON-PHL-20-01171/2020\|EPI_ISL_548793 | .................... | .................... | .................... | .................... | .................... | .................... |
| I | 3068 | hCoV-19/Canada/ON-PHL-20-01172/2020\|EPI_ISL_548794 | .................... | .................... | .................... | .................... | .................... | .................... |
| I | 3069 | hCoV-19/Canada/ON-PHL-20-01173/2020\|EPI_ISL_548795 | .................... | .................... | .................... | .................... | .................... | .................... |
| I | 3070 | hCoV-19/Canada/ON-PHL-20-01174/2020\|EPI_ISL_548796 | .................... | .................... | .................... | .................... | .................... | .................... |
| I | 3071 | hCoV-19/Canada/ON-PHL-20-01176/2020\|EPI_ISL_548798 | .................... | .................... | .................... | .................... | .................... | .................... |
| I | 3072 | hCoV-19/Canada/ON-PHL-20-01177/2020\|EPI_ISL_548799 | .................... | .................... | .................... | .................... | .................... | .................... |
| I | 3073 | hCoV-19/USA/TX-HMH-2971/2020\|EPI_ISL_543902\|2020-0 | .................... | .................... | .................... | .................... | .................... | .................... |
| I | 3074 | hCoV-19/USA/TX-HMH-2972/2020\|EPI_ISL_543903\|2020-0 | .................... | .................... | .................... | .................... | .................... | .................... |
| I | 3075 | hCoV-19/USA/TX-HMH-2973/2020\|EPI_ISL_543904\|2020-0 | .................... | .................... | .................... | .................... | .................... | .................... |
| I | 3076 | hCoV-19/USA/TX-HMH-2976/2020\|EPI_ISL_543906\|2020-0 | .................... | .................... | .................... | .................... | .................... | .................... |
| I | 3077 | hCoV-19/USA/TX-HMH-2979/2020\|EPI_ISL_543908\|2020-0 | .................... | .................... | .................... | .................... | .................... | .................... |
| I | 3080 | hCoV-19/USA/TX-HMH-2987/2020\|EPI_ISL_543914\|2020-0 | .................... | .................... | .................... | .................... | .................... | .................... |
| I | 3081 | hCoV-19/USA/TX-HMH-2991/2020\|EPI_ISL_543915\|2020-0 | .................... | .................... | .................... | .................... | .................... | .................... |
| I | 3083 | hCoV-19/USA/TX-HMH-2993/2020\|EPI_ISL_543917\|2020-0 | .................... | .................... | .................... | .................... | .................... | .................... |
| I | 3084 | hCoV-19/USA/TX-HMH-2995/2020\|EPI_ISL_543919\|2020-0 | .................... | .................... | .................... | .................... | .................... | .................... |
| I | 3085 | hCoV-19/USA/TX-HMH-2996/2020\|EPI_ISL_543920\|2020-0 | .................... | ......A............. | .................... | .................... | .................... | .................... |
| I | 3086 | hCoV-19/USA/TX-HMH-2998/2020\|EPI_ISL_543921\|2020-0 | .................... | .................... | .................... | .................... | .................... | .................... |
| I | 3087 | hCoV-19/USA/TX-HMH-3001/2020\|EPI_ISL_543923\|2020-0 | .................... | .................... | .................... | .................... | .................... | .................... |
| I | 3088 | hCoV-19/USA/TX-HMH-3040/2020\|EPI_ISL_543952\|2020-0 | .................... | .................... | .................... | .................... | .................... | .................... |
| I | 3089 | hCoV-19/USA/TX-HMH-3046/2020\|EPI_ISL_543955\|2020-0 | .................... | .................... | .................... | .................... | .................... | .......N............ |
| I | 3091 | hCoV-19/USA/TX-HMH-3051/2020\|EPI_ISL_543957\|2020-0 | .................... | .................... | .................... | .................... | .................... | .................... |
| I | 3093 | hCoV-19/USA/TX-HMH-3055/2020\|EPI_ISL_543961\|2020-0 | .................... | .................... | .................... | .................... | .................... | .................... |
| I | 3094 | hCoV-19/USA/TX-HMH-3056/2020\|EPI_ISL_543962\|2020-0 | .................... | .................... | .................... | .................... | .................... | .................... |
| I | 3095 | hCoV-19/USA/TX-HMH-3057/2020\|EPI_ISL_543963\|2020-0 | .................... | ..............C..... | .................... | .................... | .................... | .................... |
| I | 3097 | hCoV-19/USA/TX-HMH-3064/2020\|EPI_ISL_543969\|2020-0 | .................... | .................... | .................... | .................... | .................... | .................... |
| I | 3098 | hCoV-19/USA/TX-HMH-3065/2020\|EPI_ISL_543970\|2020-0 | .................... | .................... | .................... | .................... | .................... | .................... |
| I | 3099 | hCoV-19/USA/TX-HMH-3067/2020\|EPI_ISL_543972\|2020-0 | .................... | .................... | .................... | .................... | .................... | .................... |
| I | 3100 | hCoV-19/USA/TX-HMH-3069/2020\|EPI_ISL_543973\|2020-0 | .................... | .................... | .................... | .................... | .................... | .................... |
| I | 3101 | hCoV-19/USA/TX-HMH-3070/2020\|EPI_ISL_543974\|2020-0 | .................... | .................... | .................... | .................... | .................... | .................... |
| I | 3102 | hCoV-19/USA/TX-HMH-3073/2020\|EPI_ISL_543977\|2020-0 | .................... | .................... | .................... | .................... | .................... | .................... |
| I | 3103 | hCoV-19/USA/TX-HMH-3074/2020\|EPI_ISL_543978\|2020-0 | .................... | .A.................. | .................... | .................... | .................... | .................... |
| I | 3104 | hCoV-19/USA/TX-HMH-3076/2020\|EPI_ISL_543979\|2020-0 | .................... | .................... | .................... | .................... | .................... | .................... |
| I | 3106 | hCoV-19/USA/TX-HMH-3081/2020\|EPI_ISL_543982\|2020-0 | .................... | .................... | .................... | .................... | .................... | .................... |
| I | 3107 | hCoV-19/USA/TX-HMH-3082/2020\|EPI_ISL_543983\|2020-0 | .................... | .................... | .................... | .................... | .................... | .................... |
| I | 3108 | hCoV-19/USA/TX-HMH-3083/2020\|EPI_ISL_543984\|2020-0 | .................... | .................... | .................... | .................... | .................... | .................... |
| I | 3109 | hCoV-19/USA/TX-HMH-3084/2020\|EPI_ISL_543985\|2020-0 | .................... | .................... | .................... | .................... | .................... | .................... |
| I | 3110 | hCoV-19/USA/TX-HMH-3086/2020\|EPI_ISL_543987\|2020-0 | .................... | .................... | .................... | .................... | .................... | .................... |
| I | 3111 | hCoV-19/USA/TX-HMH-3092/2020\|EPI_ISL_543989\|2020-0 | .................... | .................... | .................... | .................... | .................... | .................... |

| | | | | | | | | |
|---|---|---|---|---|---|---|---|---|
| I | 3112 | hCoV-19/USA/TX-HMH-3093/2020\|EPI_ISL_543990\|2020-0 | ···················· | ···················· | ····················· | ···················· | ···················· | ···················· |
| I | 3113 | hCoV-19/USA/TX-HMH-3094/2020\|EPI_ISL_543991\|2020-0 | ···················· | ···················· | ····················· | ···················· | ···················· | ···················· |
| I | 3114 | hCoV-19/USA/TX-HMH-3095/2020\|EPI_ISL_543992\|2020-0 | ···················· | ···················· | ····················· | ···················· | ···················· | ···················· |
| I | 3115 | hCoV-19/USA/TX-HMH-3099/2020\|EPI_ISL_543993\|2020-0 | ···················· | ···················· | ····················· | ···················· | ···················· | ···················· |
| I | 3117 | hCoV-19/USA/TX-HMH-3110/2020\|EPI_ISL_543998\|2020-0 | ···················· | ···················· | ····················· | ···················· | ···················· | ···················· |
| I | 3118 | hCoV-19/USA/TX-HMH-3111/2020\|EPI_ISL_543999\|2020-0 | ···················· | ···················· | ····················· | ···················· | ···················· | ···················· |
| I | 3121 | hCoV-19/USA/TX-HMH-4768/2020\|EPI_ISL_544318\|2020-0 | ···················· | ···················· | ····················· | ···················· | ···················· | ···················· |
| I | 3127 | hCoV-19/USA/TX-HMH-4780/2020\|EPI_ISL_544325\|2020-0 | ···················· | ···················· | ····················· | ···················· | ···················· | ···················· |
| I | 3132 | hCoV-19/USA/TX-HMH-479/2020\|EPI_ISL_544332\|2020-04 | ···················· | ···················· | ····················· | ···················· | ···················· | ···················· |
| I | 3134 | hCoV-19/USA/TX-HMH-4791/2020\|EPI_ISL_544334\|2020-0 | ···················· | ···················· | ····················· | ···················· | ···················· | NN·················· |
| I | 3137 | hCoV-19/USA/TX-HMH-480/2020\|EPI_ISL_544339\|2020-04 | ···················· | ···················· | ····················· | ···················· | ···················· | ···················· |
| I | 3140 | hCoV-19/USA/TX-HMH-481/2020\|EPI_ISL_544345\|2020-04 | ···················· | ···················· | ····················· | ···················· | ···················· | ···················· |
| I | 3144 | hCoV-19/USA/TX-HMH-4817/2020\|EPI_ISL_544351\|2020-0 | ···················· | ·············A······ | ····················· | ···················· | ···················· | ···················· |
| I | 3145 | hCoV-19/USA/TX-HMH-4818/2020\|EPI_ISL_544352\|2020-0 | ···················· | ···················· | ····················· | ···················· | ···················· | ···················· |
| I | 3146 | hCoV-19/USA/TX-HMH-482/2020\|EPI_ISL_544353\|2020-04 | ···················· | ···················· | ····················· | ···················· | ···················· | ···················· |
| I | 3147 | hCoV-19/USA/TX-HMH-4820/2020\|EPI_ISL_544354\|2020-0 | ···················· | ···················· | ····················· | ···················· | ···················· | ···················· |
| I | 3148 | hCoV-19/USA/TX-HMH-4821/2020\|EPI_ISL_544355\|2020-0 | ···················· | ···················· | ····················· | ···················· | ···················· | ···················· |
| I | 3149 | hCoV-19/USA/TX-HMH-4822/2020\|EPI_ISL_544356\|2020-0 | ···················· | ···················· | ····················· | ···················· | ···················· | ···················· |
| I | 3150 | hCoV-19/USA/TX-HMH-4825/2020\|EPI_ISL_544357\|2020-0 | ···················· | ···················· | ····················· | ···················· | ···················· | ···················· |
| I | 3151 | hCoV-19/USA/TX-HMH-4827/2020\|EPI_ISL_544358\|2020-0 | ···················· | ···················· | ····················· | ···················· | ···················· | NN·················· |
| I | 3152 | hCoV-19/USA/TX-HMH-4829/2020\|EPI_ISL_544359\|2020-0 | ···················· | ···················· | ····················· | ···················· | ···················· | ···················· |
| I | 3153 | hCoV-19/USA/TX-HMH-483/2020\|EPI_ISL_544360\|2020-04 | ···················· | ···················· | ····················· | ···················· | ···················· | ···················· |
| I | 3154 | hCoV-19/USA/TX-HMH-4831/2020\|EPI_ISL_544362\|2020-0 | ···················· | ···················· | ····················· | ···················· | ···················· | ···················· |
| I | 3156 | hCoV-19/USA/TX-HMH-4834/2020\|EPI_ISL_544364\|2020-0 | ···················· | ·············A······ | ····················· | ···················· | ···N················ | N··················· |
| I | 3157 | hCoV-19/USA/TX-HMH-4836/2020\|EPI_ISL_544365\|2020-0 | ···················· | ···················· | ····················· | ···················· | ···················· | ···················· |
| I | 3158 | hCoV-19/USA/TX-HMH-4837/2020\|EPI_ISL_544366\|2020-0 | ···················· | ···················· | ····················· | ···················· | ···················· | ···················· |
| I | 3159 | hCoV-19/USA/TX-HMH-4839/2020\|EPI_ISL_544367\|2020-0 | ···················· | ···················· | ····················· | ···················· | ···················· | ···················· |
| I | 3160 | hCoV-19/USA/TX-HMH-484/2020\|EPI_ISL_544368\|2020-04 | ···················· | ···················· | ····················· | ···················· | ···················· | ···················· |
| I | 3161 | hCoV-19/USA/TX-HMH-4841/2020\|EPI_ISL_544369\|2020-0 | ···················· | ···················· | ····················· | ···················· | ···················· | ···················· |
| I | 3164 | hCoV-19/USA/TX-HMH-7811/2020\|EPI_ISL_545646\|2020-0 | ···················· | ···················· | ····················· | ···················· | ···················· | ···················· |
| I | 3165 | hCoV-19/USA/TX-HMH-7819/2020\|EPI_ISL_545647\|2020-0 | ···················· | ·············A······ | ····················· | ···················· | ···················· | ···················· |
| I | 3166 | hCoV-19/USA/TX-HMH-7822/2020\|EPI_ISL_545650\|2020-0 | ···················· | ···················· | ····················· | ···················· | ···················· | ···················· |
| I | 3167 | hCoV-19/USA/TX-HMH-7827/2020\|EPI_ISL_545651\|2020-0 | ···················· | ···················· | ····················· | ···················· | ···················· | ···················· |
| I | 3168 | hCoV-19/USA/TX-HMH-783/2020\|EPI_ISL_545653\|2020-04 | ···················· | ···················· | ····················· | ···················· | ···················· | ···················· |
| I | 3169 | hCoV-19/USA/TX-HMH-7834/2020\|EPI_ISL_545656\|2020-0 | ···················· | ··········A········· | ····················· | ···················· | ···················· | ···················· |
| I | 3170 | hCoV-19/USA/TX-HMH-7839/2020\|EPI_ISL_545658\|2020-0 | ···················· | ···················· | ····················· | ···················· | ···················· | ···················· |
| I | 3171 | hCoV-19/USA/TX-HMH-784/2020\|EPI_ISL_545659\|2020-04 | ···················· | ···················· | ····················· | ···················· | ····T··············· | ···················· |
| I | 3172 | hCoV-19/USA/TX-HMH-7841/2020\|EPI_ISL_545660\|2020-0 | ···················· | ···················· | ····················· | ···················· | ···················· | ···················· |
| I | 3173 | hCoV-19/USA/TX-HMH-7842/2020\|EPI_ISL_545661\|2020-0 | ···················· | ···················· | ····················· | ···················· | ···················· | ···················· |
| I | 3174 | hCoV-19/USA/TX-HMH-7843/2020\|EPI_ISL_545662\|2020-0 | ···················· | ···················· | ····················· | ···················· | ···················· | ···················· |
| I | 3175 | hCoV-19/USA/TX-HMH-7845/2020\|EPI_ISL_545663\|2020-0 | ···················· | ···················· | ····················· | ···················· | ···················· | ···················· |
| I | 3176 | hCoV-19/USA/TX-HMH-7846/2020\|EPI_ISL_545664\|2020-0 | ···················· | ···················· | ····················· | ···················· | ···················· | ···················· |
| I | 3177 | hCoV-19/USA/TX-HMH-7854/2020\|EPI_ISL_545666\|2020-0 | ···················· | ···················· | ····················· | ···················· | ···················· | ···················· |
| I | 3178 | hCoV-19/USA/TX-HMH-7859/2020\|EPI_ISL_545667\|2020-0 | ···················· | ······A············· | ····················· | ···················· | ···················· | ···················· |
| I | 3179 | hCoV-19/USA/TX-HMH-786/2020\|EPI_ISL_545668\|2020-04 | ···················· | ···················· | ····················· | ···················· | ···················· | ···················· |
| I | 3180 | hCoV-19/USA/TX-HMH-7860/2020\|EPI_ISL_545669\|2020-0 | ···················· | ·············A······ | ····················· | ···················· | ···················· | ···················· |
| I | 3181 | hCoV-19/USA/TX-HMH-7864/2020\|EPI_ISL_545670\|2020-0 | ···················· | ···················· | ····················· | ···················· | ···················· | ···················· |
| I | 3182 | hCoV-19/USA/TX-HMH-7868/2020\|EPI_ISL_545671\|2020-0 | ···················· | ·············A······ | ····················· | ···················· | ···················· | ···················· |
| I | 3183 | hCoV-19/USA/TX-HMH-7869/2020\|EPI_ISL_545672\|2020-0 | ···················· | ·············A······ | ····················· | ···················· | ···················· | ···················· |
| I | 3184 | hCoV-19/USA/TX-HMH-7870/2020\|EPI_ISL_545673\|2020-0 | ···················· | ···················· | ····················· | ···················· | ···················· | ···················· |
| I | 3186 | hCoV-19/USA/TX-HMH-7873/2020\|EPI_ISL_545675\|2020-0 | ···················· | ···················· | ····················· | ···················· | ···················· | ···················· |
| I | 3187 | hCoV-19/USA/TX-HMH-7874/2020\|EPI_ISL_545676\|2020-0 | ···················· | ···················· | ····················· | ···················· | ···················· | ···················· |
| I | 3188 | hCoV-19/USA/TX-HMH-7876/2020\|EPI_ISL_545677\|2020-0 | ···················· | ···················· | ····················· | ···················· | ···················· | ···················· |
| I | 3189 | hCoV-19/USA/TX-HMH-7878/2020\|EPI_ISL_545678\|2020-0 | ···················· | ···················· | ····················· | ···················· | ···················· | ···················· |
| I | 3190 | hCoV-19/USA/TX-HMH-7881/2020\|EPI_ISL_545679\|2020-0 | ···················· | ···················· | ····················· | ···················· | ···················· | ···················· |
| I | 3191 | hCoV-19/USA/TX-HMH-7883/2020\|EPI_ISL_545680\|2020-0 | ···················· | ·············A······ | ····················· | ···················· | ···················· | ···················· |
| I | 3192 | hCoV-19/USA/TX-HMH-7885/2020\|EPI_ISL_545681\|2020-0 | ···················· | ···················· | ····················· | ···················· | ···················· | ···················· |
| I | 3193 | hCoV-19/USA/TX-HMH-7886/2020\|EPI_ISL_545682\|2020-0 | ···················· | ···················· | ····················· | ···················· | ···················· | ···················· |
| I | 3195 | hCoV-19/USA/TX-HMH-7888/2020\|EPI_ISL_545684\|2020-0 | ···················· | ···················· | ····················· | ···················· | ···················· | ···················· |
| I | 3196 | hCoV-19/USA/TX-HMH-7889/2020\|EPI_ISL_545685\|2020-0 | ···················· | ···················· | ····················· | ···················· | ···················· | ···················· |
| I | 3198 | hCoV-19/USA/TX-HMH-7895/2020\|EPI_ISL_545687\|2020-0 | ···················· | ···················· | ····················· | ···················· | ···················· | ···················· |
| I | 3200 | hCoV-19/USA/TX-HMH-7897/2020\|EPI_ISL_545689\|2020-0 | ···················· | ···················· | ····················· | ···················· | ···················· | ···················· |
| I | 3201 | hCoV-19/USA/TX-HMH-790/2020\|EPI_ISL_545691\|2020-04 | ···················· | ···················· | ····················· | ···················· | ···················· | ···················· |
| I | 3202 | hCoV-19/USA/TX-HMH-7900/2020\|EPI_ISL_545692\|2020-0 | ···················· | ····T··············· | ····················· | ···················· | ···················· | ···················· |
| I | 3205 | hCoV-19/USA/TX-HMH-7904/2020\|EPI_ISL_545695\|2020-0 | ···················· | ···················· | ····················· | ···················· | ···················· | ···················· |
| I | 3207 | hCoV-19/USA/TX-HMH-7907/2020\|EPI_ISL_545697\|2020-0 | ···················· | ···················· | ····················· | ···················· | ···················· | ···················· |
| I | 3208 | hCoV-19/USA/TX-HMH-7908/2020\|EPI_ISL_545698\|2020-0 | ···················· | ····T··············· | ····················· | ···················· | ···················· | ···················· |
| I | 3209 | hCoV-19/USA/TX-HMH-7910/2020\|EPI_ISL_545700\|2020-0 | ···················· | ······A············· | ····················· | ···················· | ···················· | ···················· |
| I | 3215 | hCoV-19/USA/TX-HMH-792/2020\|EPI_ISL_545707\|2020-03 | ···················· | ···················· | ····················· | ···················· | ···················· | ···················· |
| I | 3216 | hCoV-19/USA/TX-HMH-7920/2020\|EPI_ISL_545708\|2020-0 | ···················· | ·············A······ | ····················· | ···················· | ···················· | ···················· |
| I | 3220 | hCoV-19/USA/TX-HMH-793/2020\|EPI_ISL_545714\|2020-03 | ···················· | ···················· | ····················· | ···················· | ···················· | ···················· |
| I | 3225 | hCoV-19/USA/TX-HMH-7935/2020\|EPI_ISL_545719\|2020-0 | ···················· | ···················· | ····················· | ···················· | ···················· | ···················· |
| I | 3226 | hCoV-19/USA/TX-HMH-7936/2020\|EPI_ISL_545720\|2020-0 | ···················· | ·············A······ | ····················· | ···················· | ···················· | ···················· |
| I | 3227 | hCoV-19/USA/TX-HMH-7937/2020\|EPI_ISL_545721\|2020-0 | ···················· | ···················· | ····················· | ···················· | ···················· | NN·················· |
| I | 3228 | hCoV-19/USA/TX-HMH-7938/2020\|EPI_ISL_545722\|2020-0 | ···················· | ···················· | ····················· | ···················· | ···················· | ···················· |
| I | 3229 | hCoV-19/USA/TX-HMH-7939/2020\|EPI_ISL_545723\|2020-0 | ···················· | ···················· | ····················· | ···················· | ··············N····· | NN·················· |
| I | 3230 | hCoV-19/USA/TX-HMH-794/2020\|EPI_ISL_545724\|2020-04 | ···················· | ···················· | ····················· | ···················· | ···················· | ···················· |
| I | 3233 | hCoV-19/USA/TX-HMH-7945/2020\|EPI_ISL_545728\|2020-0 | ···················· | ···················· | ····················· | ···················· | ···················· | ···················· |
| I | 3236 | hCoV-19/USA/TX-HMH-795/2020\|EPI_ISL_545732\|2020-04 | ···················· | ···················· | ····················· | ···················· | ···················· | ···················· |
| I | 3243 | hCoV-19/USA/TX-HMH-6666/2020\|EPI_ISL_546157\|2020-0 | ···················· | ···················· | ····················· | ···················· | ···················· | ···················· |

| | | | | | | | | |
|---|---|---|---|---|---|---|---|---|
| I | 3255 | hCoV-19/USA/TX-HMH-6699/2020\|EPI_ISL_546178\|2020-0 | ···················· | ···················· | ···················· | ···················· | ···················· | ···················· |
| I | 3256 | hCoV-19/USA/TX-HMH-6700/2020\|EPI_ISL_546179\|2020-0 | ···················· | ···················· | ···················· | ···················· | ···················· | ···················· |
| I | 3258 | hCoV-19/USA/TX-HMH-6702/2020\|EPI_ISL_546181\|2020-0 | ···················· | ···················· | ···················· | ···················· | ···················· | ···················· |
| I | 3259 | hCoV-19/USA/TX-HMH-6704/2020\|EPI_ISL_546183\|2020-0 | ···················· | ···················· | ···················· | ···················· | ···················· | ···················· |
| I | 3260 | hCoV-19/USA/TX-HMH-6707/2020\|EPI_ISL_546184\|2020-0 | ···················· | ···················· | ···················· | ···················· | ···················· | ···················· |
| I | 3262 | hCoV-19/USA/TX-HMH-6714/2020\|EPI_ISL_546188\|2020-0 | ···················· | ···················· | ···················· | ···················· | ···················· | ···················· |
| I | 3263 | hCoV-19/USA/TX-HMH-6716/2020\|EPI_ISL_546190\|2020-0 | ···················· | ···················· | ···················· | ···················· | ···················· | ···················· |
| I | 3266 | hCoV-19/USA/TX-HMH-6723/2020\|EPI_ISL_546193\|2020-0 | ···················· | ···················· | ···················· | ···················· | ···················· | ···················· |
| I | 3267 | hCoV-19/USA/TX-HMH-6724/2020\|EPI_ISL_546194\|2020-0 | ···················· | ···················· | ···················· | ···················· | ···················· | ···················· |
| I | 3269 | hCoV-19/USA/TX-HMH-6726/2020\|EPI_ISL_546196\|2020-0 | ···················· | ···················· | ···················· | ···················· | ···················· | ···················· |
| I | 3270 | hCoV-19/USA/TX-HMH-6727/2020\|EPI_ISL_546197\|2020-0 | ···················· | ···················· | ···················· | ···················· | ···················· | ···················· |
| I | 3274 | hCoV-19/USA/TX-HMH-5738/2020\|EPI_ISL_546979\|2020-0 | ···················· | ···················· | ···················· | ···················· | ···················· | ···················· |
| I | 3275 | hCoV-19/USA/TX-HMH-5740/2020\|EPI_ISL_546980\|2020-0 | ···················· | ···················· | ···················· | ···················· | ···················· | ···················· |
| I | 3277 | hCoV-19/USA/TX-HMH-5743/2020\|EPI_ISL_546983\|2020-0 | ···················· | ···················· | ···················· | ···················· | ···················· | ···················· |
| I | 3278 | hCoV-19/USA/TX-HMH-5745/2020\|EPI_ISL_546984\|2020-0 | ···················· | ·············A······ | ···················· | ···················· | ···················· | ···················· |
| I | 3280 | hCoV-19/USA/TX-HMH-5752/2020\|EPI_ISL_546988\|2020-0 | ···················· | ···················· | ···················· | ···················· | ···················· | ···················· |
| I | 3281 | hCoV-19/USA/TX-HMH-5753/2020\|EPI_ISL_546989\|2020-0 | ···················· | ···················· | ···················· | ···················· | ···················· | ···················· |
| I | 3282 | hCoV-19/USA/TX-HMH-5754/2020\|EPI_ISL_546990\|2020-0 | ···················· | ···················· | ···················· | ···················· | ···················· | ···················· |
| I | 3283 | hCoV-19/USA/TX-HMH-5755/2020\|EPI_ISL_546991\|2020-0 | ···················· | ···················· | ···················· | ···················· | ···················· | ···················· |
| I | 3284 | hCoV-19/USA/TX-HMH-5756/2020\|EPI_ISL_546992\|2020-0 | ···················· | ···················· | ···················· | ···················· | ···················· | ···················· |
| I | 3285 | hCoV-19/USA/TX-HMH-5759/2020\|EPI_ISL_546995\|2020-0 | ···················· | ···················· | ···················· | ···················· | ···················· | ···················· |
| I | 3287 | hCoV-19/USA/TX-HMH-5763/2020\|EPI_ISL_546997\|2020-0 | ···················· | ···················· | ···················· | ···················· | ···················· | ···················· |
| I | 3288 | hCoV-19/USA/TX-HMH-5767/2020\|EPI_ISL_546999\|2020-0 | ···················· | ···················· | ···················· | ···················· | ···················· | ···················· |
| I | 3289 | hCoV-19/Netherlands/NB-RIVM-10048/2020\|EPI_ISL_547 | ···················· | ···················· | ···················· | ···················· | ···················· | ···················· |
| I | 3290 | hCoV-19/Netherlands/NB-RIVM-10049/2020\|EPI_ISL_547 | ···················· | ···················· | ···················· | ···················· | ···················· | ···················· |
| I | 3291 | hCoV-19/Netherlands/NB-RIVM-10052/2020\|EPI_ISL_547 | ···················· | ···················· | ···················· | ···················· | ···················· | ···················· |
| I | 3292 | hCoV-19/Netherlands/NB-RIVM-10053/2020\|EPI_ISL_547 | ···················· | ···················· | ···················· | ···················· | ···················· | ···················· |
| I | 3293 | hCoV-19/Netherlands/NB-RIVM-10056/2020\|EPI_ISL_547 | ···················· | ···················· | ···················· | ···················· | ···················· | ···················· |
| I | 3294 | hCoV-19/Netherlands/NB-RIVM-10057/2020\|EPI_ISL_547 | ···················· | ···················· | ···················· | ···················· | ···················· | ···················· |
| I | 3295 | hCoV-19/Netherlands/NB-RIVM-10069/2020\|EPI_ISL_547 | ···················· | ···················· | ···················· | ···················· | ···················· | ···················· |
| I | 3296 | hCoV-19/Netherlands/NB-RIVM-10078/2020\|EPI_ISL_547 | ···················· | ···················· | ···················· | ···················· | ···················· | ···················· |
| I | 3297 | hCoV-19/Netherlands/NB-RIVM-10085/2020\|EPI_ISL_547 | ···················· | ···················· | ···················· | ···················· | ···················· | ···················· |
| I | 3298 | hCoV-19/Netherlands/NB-RIVM-10086/2020\|EPI_ISL_547 | ···················· | ···················· | ···················· | ···················· | ···················· | ···················· |
| I | 3299 | hCoV-19/Netherlands/NB-RIVM-10087/2020\|EPI_ISL_547 | ···················· | ···················· | ···················· | ···················· | ···················· | ···················· |
| I | 3300 | hCoV-19/Netherlands/NH-RIVM-10047/2020\|EPI_ISL_547 | ···················· | ···················· | ···················· | ···················· | ···················· | ···················· |
| I | 3301 | hCoV-19/Netherlands/NH-RIVM-10051/2020\|EPI_ISL_547 | ···················· | ···················· | ···················· | ···················· | ···················· | ···················· |
| I | 3302 | hCoV-19/Netherlands/NH-RIVM-10058/2020\|EPI_ISL_547 | ···················· | ···················· | ···················· | ···················· | ···················· | ···················· |
| I | 3303 | hCoV-19/Netherlands/NH-RIVM-10061/2020\|EPI_ISL_547 | ···················· | ···················· | ···················· | ···················· | ···················· | ···················· |
| I | 3304 | hCoV-19/Netherlands/NH-RIVM-10062/2020\|EPI_ISL_547 | ···················· | ···················· | ···················· | ···················· | ···················· | ···················· |
| I | 3305 | hCoV-19/Netherlands/NH-RIVM-10067/2020\|EPI_ISL_547 | ···················· | ··········A········· | ···················· | ···················· | ···················· | ···················· |
| I | 3306 | hCoV-19/Netherlands/NH-RIVM-10070/2020\|EPI_ISL_547 | ···················· | ··········A········· | ···················· | ···················· | ···················· | ···················· |
| I | 3307 | hCoV-19/Netherlands/NH-RIVM-10071/2020\|EPI_ISL_547 | ···················· | ··········A········· | ···················· | ···················· | ···················· | ···················· |
| I | 3308 | hCoV-19/Netherlands/NH-RIVM-10073/2020\|EPI_ISL_547 | ···················· | ···················· | ···················· | ···················· | ···················· | ···················· |
| I | 3309 | hCoV-19/Canada/ON-PHL-20-00019/2020\|EPI_ISL_548700 | ···················· | ···················· | ···················· | ···················· | ···················· | ···················· |
| I | 3310 | hCoV-19/Canada/ON-PHL-20-00020/2020\|EPI_ISL_548701 | ···················· | ···················· | ···················· | ···················· | ···················· | ···················· |
| I | 3311 | hCoV-19/Canada/ON-PHL-20-00021/2020\|EPI_ISL_548702 | ···················· | ···················· | ···················· | ···················· | ···················· | ···················· |
| I | 3312 | hCoV-19/Canada/ON-PHL-20-00022/2020\|EPI_ISL_548703 | ···················· | ···················· | ···················· | ···················· | ···················· | ···················· |
| I | 3313 | hCoV-19/Canada/ON-PHL-20-00023/2020\|EPI_ISL_548704 | ···················· | ···················· | ···················· | ···················· | ···················· | ···················· |
| I | 3314 | hCoV-19/Canada/ON-PHL-20-00024/2020\|EPI_ISL_548705 | ···················· | ···················· | ···················· | ···················· | ···················· | ···················· |
| I | 3315 | hCoV-19/Canada/ON-PHL-20-00027/2020\|EPI_ISL_548708 | ···················· | ···················· | ···················· | ···················· | ···················· | ···················· |
| I | 3316 | hCoV-19/Canada/ON-PHL-20-00028/2020\|EPI_ISL_548709 | ···················· | ···················· | ···················· | ···················· | ···················· | ···················· |
| I | 3317 | hCoV-19/Canada/ON-PHL-20-00030/2020\|EPI_ISL_548711 | ···················· | ···················· | ···················· | ···················· | ···················· | ···················· |
| I | 3318 | hCoV-19/Canada/ON-PHL-20-00031/2020\|EPI_ISL_548712 | ···················· | ···················· | ···················· | ···················· | ···················· | ···················· |
| I | 3319 | hCoV-19/Canada/ON-PHL-20-00032/2020\|EPI_ISL_548713 | ···················· | ···················· | ···················· | ···················· | ···················· | ···················· |
| I | 3320 | hCoV-19/Canada/ON-PHL-20-00033/2020\|EPI_ISL_548714 | ···················· | ···················· | ···················· | ···················· | ···················· | ···················· |
| I | 3321 | hCoV-19/Canada/ON-PHL-20-00037/2020\|EPI_ISL_548716 | ···················· | ···················· | ···················· | ···················· | ···················· | ···················· |
| I | 3322 | hCoV-19/Canada/ON-PHL-20-00038/2020\|EPI_ISL_548717 | ···················· | ···················· | ···················· | ···················· | ···················· | ···················· |
| I | 3323 | hCoV-19/Canada/ON-PHL-20-00039/2020\|EPI_ISL_548718 | ···················· | ···················· | ···················· | ···················· | ···················· | ···················· |
| I | 3324 | hCoV-19/Canada/ON-PHL-20-00042/2020\|EPI_ISL_548720 | ···················· | ···················· | ···················· | ···················· | ···················· | ···················· |
| I | 3325 | hCoV-19/Canada/ON-PHL-20-00043/2020\|EPI_ISL_548721 | ···················· | ···················· | ···················· | ···················· | ···················· | ···················· |
| I | 3326 | hCoV-19/Canada/ON-PHL-20-00044/2020\|EPI_ISL_548722 | ···················· | ···················· | ···················· | ···················· | ···················· | ···················· |
| I | 3327 | hCoV-19/Canada/ON-PHL-20-00045/2020\|EPI_ISL_548723 | ···················· | ···················· | ···················· | ···················· | ···················· | ···················· |
| I | 3328 | hCoV-19/Canada/ON-PHL-20-00046/2020\|EPI_ISL_548724 | ···················· | ···················· | ···················· | ···················· | ···················· | ···················· |
| I | 3330 | hCoV-19/USA/TX-HMH-2824/2020\|EPI_ISL_543799\|2020-0 | ···················· | ···················· | ···················· | ···················· | ···················· | ···················· |
| I | 3331 | hCoV-19/USA/TX-HMH-3003/2020\|EPI_ISL_543925\|2020-0 | ···················· | ···················· | ···················· | ···················· | ···················· | ···················· |
| I | 3334 | hCoV-19/USA/TX-HMH-3009/2020\|EPI_ISL_543929\|2020-0 | ···················· | ···················· | ···················· | ···················· | ···················· | ···················· |
| I | 3335 | hCoV-19/USA/TX-HMH-3010/2020\|EPI_ISL_543930\|2020-0 | ···················· | ···················· | ···················· | ···················· | ···················· | ···················· |
| I | 3336 | hCoV-19/USA/TX-HMH-3011/2020\|EPI_ISL_543931\|2020-0 | ···················· | ···················· | ···················· | ···················· | ···················· | ···················· |
| I | 3337 | hCoV-19/USA/TX-HMH-3012/2020\|EPI_ISL_543932\|2020-0 | ···················· | ···················· | ···················· | ···················· | ··············N····· | N··N················ |
| I | 3338 | hCoV-19/USA/TX-HMH-3013/2020\|EPI_ISL_543933\|2020-0 | ···················· | ···················· | ···················· | ···················· | ···················· | ···················· |
| I | 3339 | hCoV-19/USA/TX-HMH-3015/2020\|EPI_ISL_543935\|2020-0 | ···················· | ···················· | ···················· | ···················· | ···················· | ···················· |
| I | 3340 | hCoV-19/USA/TX-HMH-3016/2020\|EPI_ISL_543936\|2020-0 | ···················· | ···················· | ···················· | ···················· | ···················· | ···················· |
| I | 3342 | hCoV-19/USA/TX-HMH-3022/2020\|EPI_ISL_543940\|2020-0 | ···················· | ···················· | ···················· | ···················· | ···················· | ···················· |
| I | 3343 | hCoV-19/USA/TX-HMH-3025/2020\|EPI_ISL_543943\|2020-0 | ···················· | ···················· | ···················· | ···················· | ···················· | ···················· |
| I | 3344 | hCoV-19/USA/TX-HMH-3026/2020\|EPI_ISL_543944\|2020-0 | ···················· | ···················· | ···················· | ···················· | ···················· | ···················· |
| I | 3345 | hCoV-19/USA/TX-HMH-3032/2020\|EPI_ISL_543946\|2020-0 | ···················· | ···················· | ···················· | ···················· | ···················· | ···················· |
| I | 3346 | hCoV-19/USA/TX-HMH-3033/2020\|EPI_ISL_543947\|2020-0 | ···················· | ···················· | ···················· | ···················· | ···················· | ···················· |
| I | 3347 | hCoV-19/USA/TX-HMH-3034/2020\|EPI_ISL_543948\|2020-0 | ···················· | ···················· | ···················· | ···················· | ···················· | ···N················ |
| I | 3348 | hCoV-19/USA/TX-HMH-3038/2020\|EPI_ISL_543950\|2020-0 | ···················· | ···················· | ···················· | ···················· | ···················· | ···················· |
| I | 3349 | hCoV-19/USA/TX-HMH-4678/2020\|EPI_ISL_544249\|2020-0 | ···················· | ···················· | ···················· | ···················· | ···················· | ···················· |

| | | | | | | | | |
|---|---|---|---|---|---|---|---|---|
| I | 3350 hCoV-19/USA/TX-HMH-468/2020\|EPI_ISL_544251\|2020-04 | ···················· | ···················· | ····················· | ···················· | ···················· | ···················· | |
| I | 3351 hCoV-19/USA/TX-HMH-4683/2020\|EPI_ISL_544254\|2020-0 | ···················· | ···················· | ····················· | ···················· | ···················· | ···················· | |
| I | 3352 hCoV-19/USA/TX-HMH-4684/2020\|EPI_ISL_544255\|2020-0 | ···················· | ···················· | ····················· | ···················· | ···················· | ···················· | |
| I | 3354 hCoV-19/USA/TX-HMH-469/2020\|EPI_ISL_544259\|2020-04 | ···················· | ···················· | ····················· | ···················· | ···················· | ···················· | |
| I | 3356 hCoV-19/USA/TX-HMH-4694/2020\|EPI_ISL_544264\|2020-0 | ···················· | ···················· | ····················· | ···················· | ···················· | ···················· | |
| I | 3357 hCoV-19/USA/TX-HMH-4696/2020\|EPI_ISL_544265\|2020-0 | ···················· | ·············A······ | ····················· | ···················· | ···················· | ···················· | |
| I | 3359 hCoV-19/USA/TX-HMH-470/2020\|EPI_ISL_544268\|2020-04 | ···················· | ···················· | ····················· | ···················· | ···················· | ···················· | |
| I | 3361 hCoV-19/USA/TX-HMH-4703/2020\|EPI_ISL_544271\|2020-0 | ···················· | ···················· | ····················· | ···················· | ···················· | ···················· | |
| I | 3363 hCoV-19/USA/TX-HMH-4707/2020\|EPI_ISL_544275\|2020-0 | ···················· | ···················· | ····················· | ···················· | ···················· | ···A················ | |
| I | 3365 hCoV-19/USA/TX-HMH-471/2020\|EPI_ISL_544278\|2020-04 | ···················· | ···················· | ····················· | ···················· | ···················· | ···················· | |
| I | 3366 hCoV-19/USA/TX-HMH-4710/2020\|EPI_ISL_544279\|2020-0 | ···················· | ··············C····· | ····················· | ···················· | ···················· | ···················· | |
| I | 3367 hCoV-19/USA/TX-HMH-4715/2020\|EPI_ISL_544281\|2020-0 | ···················· | ···················· | ····················· | ···················· | ···················· | ···················· | |
| I | 3368 hCoV-19/USA/TX-HMH-4717/2020\|EPI_ISL_544282\|2020-0 | ···················· | ··········A········· | ····················· | ···················· | ···················· | ···················· | |
| I | 3369 hCoV-19/USA/TX-HMH-4718/2020\|EPI_ISL_544283\|2020-0 | ···················· | ···················· | ····················· | ···················· | ···················· | ···················· | |
| I | 3371 hCoV-19/USA/TX-HMH-472/2020\|EPI_ISL_544285\|2020-04 | ···················· | ···················· | ····················· | ···················· | ···················· | ···················· | |
| I | 3376 hCoV-19/USA/TX-HMH-474/2020\|EPI_ISL_544296\|2020-04 | ···················· | ···················· | ····················· | ···················· | ···················· | ···················· | |
| I | 3381 hCoV-19/USA/TX-HMH-2718/2020\|EPI_ISL_545501\|2020-0 | ···················· | ···················· | ····················· | ···················· | ···················· | ···················· | |
| I | 3382 hCoV-19/USA/TX-HMH-2720/2020\|EPI_ISL_545503\|2020-0 | ···················· | ···················· | ····················· | ···················· | ···················· | ···················· | |
| I | 3383 hCoV-19/USA/TX-HMH-2721/2020\|EPI_ISL_545504\|2020-0 | ···················· | ···················· | ····················· | ···················· | ···················· | ···················· | |
| I | 3384 hCoV-19/USA/TX-HMH-2722/2020\|EPI_ISL_545505\|2020-0 | ···················· | ···················· | ····················· | ···················· | ···················· | ···················· | |
| I | 3385 hCoV-19/USA/TX-HMH-2723/2020\|EPI_ISL_545506\|2020-0 | ···················· | ···················· | ····················· | ···················· | ···················· | ···················· | |
| I | 3386 hCoV-19/USA/TX-HMH-2724/2020\|EPI_ISL_545507\|2020-0 | ···················· | ···················· | ····················· | ···················· | ···················· | ···················· | |
| I | 3387 hCoV-19/USA/TX-HMH-2727/2020\|EPI_ISL_545509\|2020-0 | ···················· | ···················· | ····················· | ···················· | ···················· | ···················· | |
| I | 3388 hCoV-19/USA/TX-HMH-2731/2020\|EPI_ISL_545510\|2020-0 | ···················· | ···················· | ····················· | ···················· | ···················· | ···················· | |
| I | 3389 hCoV-19/USA/TX-HMH-2732/2020\|EPI_ISL_545511\|2020-0 | ···················· | ···················· | ····················· | ···················· | ···················· | ···················· | |
| I | 3390 hCoV-19/USA/TX-HMH-2733/2020\|EPI_ISL_545512\|2020-0 | ···················· | ···················· | ····················· | ···················· | ···················· | ···················· | |
| I | 3391 hCoV-19/USA/TX-HMH-2734/2020\|EPI_ISL_545513\|2020-0 | ···················· | ···················· | ····················· | ···················· | ···················· | ···················· | |
| I | 3392 hCoV-19/USA/TX-HMH-2735/2020\|EPI_ISL_545514\|2020-0 | ···················· | ···················· | ····················· | ···················· | ···················· | ···················· | |
| I | 3393 hCoV-19/USA/TX-HMH-2736/2020\|EPI_ISL_545515\|2020-0 | ···················· | ···················· | ····················· | ···················· | ···················· | ···················· | |
| I | 3394 hCoV-19/USA/TX-HMH-2737/2020\|EPI_ISL_545516\|2020-0 | ···················· | ···················· | ····················· | ···················· | N··················· | NN·····N············ | 1 |
| I | 3395 hCoV-19/USA/TX-HMH-2739/2020\|EPI_ISL_545518\|2020-0 | ···················· | ···················· | ····················· | ···················· | ···················· | ···················· | |
| I | 3396 hCoV-19/USA/TX-HMH-2740/2020\|EPI_ISL_545519\|2020-0 | ···················· | ···················· | ····················· | ···················· | ···················· | ···················· | |
| I | 3397 hCoV-19/USA/TX-HMH-2742/2020\|EPI_ISL_545520\|2020-0 | ···················· | ···················· | ····················· | ···················· | ···················· | ···················· | |
| I | 3398 hCoV-19/USA/TX-HMH-2743/2020\|EPI_ISL_545521\|2020-0 | ···················· | ···················· | ····················· | ···················· | ···················· | ···················· | |
| I | 3399 hCoV-19/USA/TX-HMH-2745/2020\|EPI_ISL_545522\|2020-0 | ···················· | ···················· | ····················· | ···················· | ···················· | ···················· | |
| I | 3400 hCoV-19/USA/TX-HMH-2746/2020\|EPI_ISL_545523\|2020-0 | ···················· | ···················· | ····················· | ···················· | ···················· | ···················· | |
| I | 3402 hCoV-19/USA/TX-HMH-2754/2020\|EPI_ISL_545530\|2020-0 | ···················· | ···················· | ····················· | ···················· | ···················· | ···················· | |
| I | 3403 hCoV-19/Czech Republic/NRL_7917/2020\|EPI_ISL_54558 | ···················· | ···················· | ····················· | ···················· | ···················· | ···················· | |
| I | 3404 hCoV-19/USA/TX-HMH-7681/2020\|EPI_ISL_545582\|2020-0 | ···················· | ·············A······ | ····················· | ···················· | ···················· | ···················· | |
| I | 3405 hCoV-19/USA/TX-HMH-7682/2020\|EPI_ISL_545583\|2020-0 | ···················· | ·A·················· | ····················· | ···················· | ··············N····· | ···················· | |
| I | 3408 hCoV-19/USA/TX-HMH-7691/2020\|EPI_ISL_545587\|2020-0 | ···················· | ···················· | ····················· | ···················· | ···················· | ···················· | |
| I | 3410 hCoV-19/USA/TX-HMH-7697/2020\|EPI_ISL_545589\|2020-0 | ···················· | ···················· | ····················· | ···················· | ···················· | ···················· | |
| I | 3411 hCoV-19/USA/TX-HMH-7702/2020\|EPI_ISL_545591\|2020-0 | ···················· | ···················· | ····················· | ···················· | ···················· | ···················· | |
| I | 3412 hCoV-19/USA/TX-HMH-7705/2020\|EPI_ISL_545593\|2020-0 | ···················· | ···················· | ····················· | ···················· | ··············N····· | ···················· | |
| I | 3413 hCoV-19/USA/TX-HMH-7707/2020\|EPI_ISL_545595\|2020-0 | ···················· | ···················· | ····················· | ···················· | ···················· | ···················· | |
| I | 3414 hCoV-19/USA/TX-HMH-7710/2020\|EPI_ISL_545596\|2020-0 | ···················· | ···················· | ····················· | ···················· | ···················· | ···················· | |
| I | 3416 hCoV-19/USA/TX-HMH-7715/2020\|EPI_ISL_545599\|2020-0 | ···················· | ·············A······ | ····················· | ···················· | ···················· | ·······N············ | |
| I | 3417 hCoV-19/USA/TX-HMH-6443/2020\|EPI_ISL_546000\|2020-0 | ···················· | ···················· | ····················· | ···················· | ···················· | ···················· | |
| I | 3421 hCoV-19/USA/TX-HMH-6452/2020\|EPI_ISL_546005\|2020-0 | ···················· | ···················· | ····················· | ···················· | ···················· | ···················· | |
| I | 3422 hCoV-19/USA/TX-HMH-6453/2020\|EPI_ISL_546006\|2020-0 | ···················· | ······A············· | ····················· | ···················· | ···················· | ···················· | |
| I | 3423 hCoV-19/USA/TX-HMH-6454/2020\|EPI_ISL_546007\|2020-0 | ···················· | ···················· | ····················· | ···················· | ···················· | ···················· | |
| I | 3424 hCoV-19/USA/TX-HMH-6455/2020\|EPI_ISL_546008\|2020-0 | ···················· | ···················· | ····················· | ···················· | ···················· | ···················· | |
| I | 3425 hCoV-19/USA/TX-HMH-6456/2020\|EPI_ISL_546009\|2020-0 | ···················· | ···················· | ····················· | ···················· | ···················· | ···················· | |
| I | 3426 hCoV-19/USA/TX-HMH-6457/2020\|EPI_ISL_546010\|2020-0 | ···················· | ···················· | ····················· | ···················· | ···················· | ···················· | |
| I | 3427 hCoV-19/USA/TX-HMH-6458/2020\|EPI_ISL_546011\|2020-0 | ···················· | ···················· | ····················· | ···················· | ···················· | ···················· | |
| I | 3429 hCoV-19/USA/TX-HMH-6461/2020\|EPI_ISL_546014\|2020-0 | ···················· | ···················· | ····················· | ···················· | ···················· | ···················· | |
| I | 3430 hCoV-19/USA/TX-HMH-6466/2020\|EPI_ISL_546017\|2020-0 | ···················· | ···················· | ····················· | ···················· | ···················· | ···················· | |
| I | 3432 hCoV-19/USA/TX-HMH-647/2020\|EPI_ISL_546019\|2020-04 | ···················· | ···················· | ····················· | ···················· | ···················· | ···················· | |
| I | 3433 hCoV-19/USA/TX-HMH-6470/2020\|EPI_ISL_546020\|2020-0 | ···················· | ···················· | ····················· | ···················· | ···················· | ···················· | |
| I | 3434 hCoV-19/USA/TX-HMH-6471/2020\|EPI_ISL_546021\|2020-0 | ···················· | ···················· | ····················· | ···················· | ···················· | ··········NN········ | |
| I | 3435 hCoV-19/USA/TX-HMH-6474/2020\|EPI_ISL_546022\|2020-0 | ···················· | ···················· | ····················· | ···················· | ···················· | ···················· | |
| I | 3437 hCoV-19/USA/TX-HMH-6479/2020\|EPI_ISL_546025\|2020-0 | ···················· | ·············A······ | ····················· | ···················· | ···················· | ···················· | |
| I | 3438 hCoV-19/USA/TX-HMH-6480/2020\|EPI_ISL_546026\|2020-0 | ···················· | ···················· | ····················· | ···················· | ···················· | ···················· | |
| I | 3439 hCoV-19/USA/TX-HMH-6481/2020\|EPI_ISL_546027\|2020-0 | ···················· | ···················· | ····················· | ···················· | ···················· | ···················· | |
| I | 3440 hCoV-19/USA/TX-HMH-6482/2020\|EPI_ISL_546028\|2020-0 | ···················· | ···················· | ····················· | ···················· | ···················· | ···················· | |
| I | 3441 hCoV-19/USA/TX-HMH-6484/2020\|EPI_ISL_546029\|2020-0 | ···················· | ···················· | ····················· | ···················· | ···················· | ···················· | |
| I | 3442 hCoV-19/USA/TX-HMH-753/2020\|EPI_ISL_546824\|2020-04 | ···················· | ···················· | ····················· | ···················· | ···················· | ···················· | |
| I | 3443 hCoV-19/USA/TX-HMH-7530/2020\|EPI_ISL_546825\|2020-0 | ···················· | ···················· | ····················· | ···················· | ···················· | ···················· | |
| I | 3445 hCoV-19/USA/TX-HMH-7532/2020\|EPI_ISL_546827\|2020-0 | ···················· | ···················· | ····················· | ···················· | ···················· | ···················· | |
| I | 3446 hCoV-19/USA/TX-HMH-7534/2020\|EPI_ISL_546829\|2020-0 | ···················· | ···················· | ····················· | ···················· | ···················· | ···················· | |
| I | 3447 hCoV-19/USA/TX-HMH-7537/2020\|EPI_ISL_546831\|2020-0 | ···················· | ···················· | ····················· | ···················· | ···················· | ···················· | |
| I | 3448 hCoV-19/USA/TX-HMH-7539/2020\|EPI_ISL_546833\|2020-0 | ···················· | ···················· | ····················· | ···················· | ···················· | ···················· | |
| I | 3449 hCoV-19/USA/TX-HMH-754/2020\|EPI_ISL_546834\|2020-04 | ···················· | ···················· | ····················· | ···················· | ···················· | ···················· | |
| I | 3450 hCoV-19/USA/TX-HMH-7542/2020\|EPI_ISL_546837\|2020-0 | ···················· | ···················· | ····················· | ···················· | ···················· | ···················· | |
| I | 3454 hCoV-19/USA/TX-HMH-7546/2020\|EPI_ISL_546841\|2020-0 | ···················· | ···················· | ····················· | ···················· | ···················· | ···················· | |
| I | 3455 hCoV-19/USA/TX-HMH-7547/2020\|EPI_ISL_546842\|2020-0 | ···················· | ···················· | ····················· | ···················· | ···················· | ···················· | |
| I | 3456 hCoV-19/USA/TX-HMH-755/2020\|EPI_ISL_546843\|2020-04 | ···················· | ···················· | ····················· | ···················· | ···················· | ···················· | |
| I | 3460 hCoV-19/USA/TX-HMH-7558/2020\|EPI_ISL_546850\|2020-0 | ···················· | ······A············· | ····················· | ···················· | ···················· | NN·····N············ | 1 |
| I | 3465 hCoV-19/USA/TX-HMH-7569/2020\|EPI_ISL_546860\|2020-0 | ···················· | ·············A······ | ····················· | ···················· | ···················· | NN·N················ | 1 |
| I | 3466 hCoV-19/USA/TX-HMH-619/2020\|EPI_ISL_547300\|2020-04 | ···················· | ···················· | ····················· | ···················· | ···················· | ···················· | |

| | | | | | | | | |
|---|---|---|---|---|---|---|---|---|
| I | 3467 | hCoV-19/USA/TX-HMH-6191/2020\|EPI_ISL_547301\|2020-0 | ···················· | ···················· | ···················· | ···················· | ···················· | ···················· |
| I | 3468 | hCoV-19/USA/TX-HMH-6192/2020\|EPI_ISL_547302\|2020-0 | ···················· | ·············A······ | ···················· | ···················· | ···················· | ···················· |
| I | 3471 | hCoV-19/USA/TX-HMH-6198/2020\|EPI_ISL_547306\|2020-0 | ···················· | ·············A······ | ···················· | ···················· | ···················· | ···················· |
| I | 3473 | hCoV-19/USA/TX-HMH-6205/2020\|EPI_ISL_547310\|2020-0 | ···················· | ···················· | ···················· | ···················· | ···················· | ···················· |
| I | 3474 | hCoV-19/USA/TX-HMH-6206/2020\|EPI_ISL_547311\|2020-0 | ···················· | ···················· | ···················· | ···················· | ···················· | ···················· |
| I | 3475 | hCoV-19/USA/TX-HMH-6207/2020\|EPI_ISL_547312\|2020-0 | ···················· | ···················· | ···················· | ···················· | ···················· | ···················· |
| I | 3476 | hCoV-19/USA/TX-HMH-6208/2020\|EPI_ISL_547313\|2020-0 | ···················· | ···················· | ···················· | ···················· | ···················· | ···················· |
| I | 3477 | hCoV-19/USA/TX-HMH-6209/2020\|EPI_ISL_547314\|2020-0 | ···················· | ···················· | ···················· | ···················· | ···················· | ···················· |
| I | 3478 | hCoV-19/USA/TX-HMH-6212/2020\|EPI_ISL_547315\|2020-0 | ···················· | ······A············· | ···················· | ···················· | ···················· | ···················· |
| I | 3479 | hCoV-19/USA/TX-HMH-6213/2020\|EPI_ISL_547316\|2020-0 | ···················· | ···················· | ···················· | ···················· | ···················· | ···················· |
| I | 3480 | hCoV-19/USA/TX-HMH-6215/2020\|EPI_ISL_547318\|2020-0 | ···················· | ···················· | ···················· | ···················· | ···················· | ···················· |
| I | 3482 | hCoV-19/USA/TX-HMH-6218/2020\|EPI_ISL_547321\|2020-0 | ···················· | ···················· | ···················· | ···················· | ··············N····· | ··N················· |
| I | 3484 | hCoV-19/USA/TX-HMH-6221/2020\|EPI_ISL_547323\|2020-0 | ···················· | ···················· | ···················· | ···················· | ···················· | ···················· |
| I | 3485 | hCoV-19/USA/TX-HMH-6223/2020\|EPI_ISL_547324\|2020-0 | ···················· | ···················· | ···················· | ···················· | ···················· | ···················· |
| I | 3488 | hCoV-19/USA/TX-HMH-6230/2020\|EPI_ISL_547328\|2020-0 | ···················· | ·············A······ | ···················· | ···················· | ···················· | ···················· |
| I | 3489 | hCoV-19/USA/TX-HMH-6234/2020\|EPI_ISL_547331\|2020-0 | ···················· | ···················· | ···················· | ···················· | ··············N····· | ···················· |
| I | 3490 | hCoV-19/USA/TX-HMH-6238/2020\|EPI_ISL_547333\|2020-0 | ···················· | ···················· | ···················· | ···················· | ···················· | ···················· |
| I | 3492 | hCoV-19/USA/TX-HMH-6242/2020\|EPI_ISL_547335\|2020-0 | ···················· | ···················· | ···················· | ···················· | ···················· | ···················· |
| I | 3493 | hCoV-19/USA/TX-HMH-6243/2020\|EPI_ISL_547336\|2020-0 | ···················· | ···················· | ···················· | ···················· | ···················· | ···················· |
| I | 3496 | hCoV-19/USA/TX-HMH-625/2020\|EPI_ISL_547340\|2020-04 | ···················· | ···················· | ···················· | ···················· | ···················· | ···················· |
| I | 3498 | hCoV-19/USA/TX-HMH-6254/2020\|EPI_ISL_547342\|2020-0 | ···················· | ···················· | ···················· | ···················· | ···················· | ···················· |
| I | 3500 | hCoV-19/USA/TX-HMH-6256/2020\|EPI_ISL_547344\|2020-0 | ···················· | ·············A······ | ···················· | ···················· | ···················· | ···················· |
| I | 3502 | hCoV-19/USA/TX-HMH-6261/2020\|EPI_ISL_547347\|2020-0 | ···················· | ···················· | ···················· | ···················· | ···················· | ···················· |
| I | 3503 | hCoV-19/USA/TX-HMH-6263/2020\|EPI_ISL_547348\|2020-0 | ···················· | ···················· | ···················· | ···················· | ···················· | ···················· |
| I | 3504 | hCoV-19/USA/TX-HMH-6265/2020\|EPI_ISL_547350\|2020-0 | ···················· | ···················· | ···················· | ···················· | ···················· | ···················· |
| I | 3505 | hCoV-19/USA/TX-HMH-6267/2020\|EPI_ISL_547352\|2020-0 | ···················· | ···················· | ···················· | ···················· | ···················· | ···················· |
| I | 3506 | hCoV-19/USA/TX-HMH-6268/2020\|EPI_ISL_547353\|2020-0 | ···················· | ···················· | ···················· | ···················· | ···················· | ···················· |
| I | 3507 | hCoV-19/USA/TX-HMH-6269/2020\|EPI_ISL_547354\|2020-0 | ···················· | ···················· | ···················· | ···················· | ···················· | ···················· |
| I | 3508 | hCoV-19/USA/TX-HMH-6274/2020\|EPI_ISL_547357\|2020-0 | ···················· | ···················· | ···················· | ···················· | ···················· | ···················· |
| I | 3509 | hCoV-19/USA/TX-HMH-6276/2020\|EPI_ISL_547359\|2020-0 | ···················· | ·A·················· | ···················· | ···················· | ···················· | ···················· |
| I | 3510 | hCoV-19/USA/CA-CZB-3997/2020\|EPI_ISL_548605\|2020-0 | ···················· | ·A·················· | ···················· | ···················· | ···················· | ···················· |
| I | 3511 | hCoV-19/USA/CA-CZB-4033/2020\|EPI_ISL_548606\|2020-0 | ···················· | ·A·················· | ···················· | ···················· | ···················· | ···················· |
| I | 3512 | hCoV-19/USA/CA-CZB-4048/2020\|EPI_ISL_548607\|2020-0 | ···················· | ·A·················· | ···················· | ···················· | ···················· | ···················· |
| I | 3513 | hCoV-19/USA/CA-CZB-4096/2020\|EPI_ISL_548608\|2020-0 | ···················· | ·A·················· | ···················· | ···················· | ···················· | ···················· |
| I | 3514 | hCoV-19/USA/CA-CZB-4087/2020\|EPI_ISL_548609\|2020-0 | ···················· | ···················· | ···················· | ···················· | ···················· | ···················· |
| I | 3515 | hCoV-19/USA/CA-CZB-4041/2020\|EPI_ISL_548610\|2020-0 | ···················· | ·A·················· | ···················· | ···················· | ···················· | ···················· |
| I | 3516 | hCoV-19/USA/CA-CZB-4073/2020\|EPI_ISL_548611\|2020-0 | ···················· | ···················· | ···················· | ···················· | ···················· | ···················· |
| I | 3517 | hCoV-19/USA/CA-CZB-4009/2020\|EPI_ISL_548612\|2020-0 | ···················· | ·A·················· | ···················· | ···················· | ···················· | ···················· |
| I | 3518 | hCoV-19/USA/CA-CZB-4083/2020\|EPI_ISL_548613\|2020-0 | ···················· | ·A·················· | ···················· | ···················· | ···················· | ···················· |
| I | 3519 | hCoV-19/USA/CA-CZB-4088/2020\|EPI_ISL_548614\|2020-0 | ···················· | ·A····A············· | ···················· | ···················· | ···················· | ···················· |
| I | 3520 | hCoV-19/USA/CA-CZB-4093/2020\|EPI_ISL_548615\|2020-0 | ···················· | ·A·················· | ···················· | ···················· | ···················· | ···················· |
| I | 3521 | hCoV-19/USA/CA-CZB-4050/2020\|EPI_ISL_548616\|2020-0 | ···················· | ···················· | ···················· | ···················· | ···················· | ···················· |
| I | 3522 | hCoV-19/USA/CA-CZB-4022/2020\|EPI_ISL_548617\|2020-0 | ···················· | ·A·················· | ···················· | ···················· | ···················· | ···················· |
| I | 3523 | hCoV-19/USA/CA-CZB-4007/2020\|EPI_ISL_548618\|2020-0 | ···················· | ·A·················· | ···················· | ···················· | ···················· | ···················· |
| I | 3524 | hCoV-19/USA/CA-CZB-4013/2020\|EPI_ISL_548619\|2020-0 | ···················· | ·A·················· | ···················· | ···················· | ···················· | ···················· |
| I | 3525 | hCoV-19/USA/CA-CZB-4089/2020\|EPI_ISL_548620\|2020-0 | ···················· | ···················· | ···················· | ···················· | ···················· | ···················· |
| I | 3526 | hCoV-19/USA/CA-CZB-4019/2020\|EPI_ISL_548621\|2020-0 | ···················· | ·A·················· | ···················· | ···················· | ···················· | ···················· |
| I | 3527 | hCoV-19/USA/CA-CZB-4042/2020\|EPI_ISL_548622\|2020-0 | ···················· | ·A·················· | ···················· | ···················· | ···················· | ···················· |
| I | 3528 | hCoV-19/USA/CA-CZB-4086/2020\|EPI_ISL_548623\|2020-0 | ···················· | ···················· | ···················· | ···················· | ···················· | ···················· |
| I | 3529 | hCoV-19/USA/CA-CZB-4011/2020\|EPI_ISL_548624\|2020-0 | ···················· | ·A·················· | ···················· | ···················· | ···················· | ···················· |
| I | 3530 | hCoV-19/USA/CA-CZB-4085/2020\|EPI_ISL_548625\|2020-0 | ···················· | ···················· | ···················· | ···················· | ···················· | ···················· |
| I | 3531 | hCoV-19/USA/CA-CZB-4101/2020\|EPI_ISL_548626\|2020-0 | ···················· | ···················· | ···················· | ···················· | ···················· | ···················· |
| I | 3532 | hCoV-19/USA/CA-CZB-4078/2020\|EPI_ISL_548627\|2020-0 | ···················· | ···················· | ···················· | ···················· | ···················· | ···················· |
| I | 3533 | hCoV-19/USA/CA-CZB-4060/2020\|EPI_ISL_548628\|2020-0 | ···················· | ···················· | ···················· | ···················· | ···················· | ···················· |
| I | 3535 | hCoV-19/USA/CA-CZB-4005/2020\|EPI_ISL_548630\|2020-0 | ···················· | ·A·················· | ···················· | ···················· | ···················· | ···················· |
| I | 3536 | hCoV-19/USA/CA-CZB-4053/2020\|EPI_ISL_548631\|2020-0 | ···················· | ·A·················· | ···················· | ···················· | ···················· | ···················· |
| I | 3537 | hCoV-19/USA/CA-CZB-3996/2020\|EPI_ISL_548632\|2020-0 | ···················· | ···················· | ···················· | ···················· | ···················· | ···················· |
| I | 3538 | hCoV-19/USA/CA-CZB-4035/2020\|EPI_ISL_548633\|2020-0 | ···················· | ·A·················· | ···················· | ···················· | ···················· | ···················· |
| I | 3539 | hCoV-19/USA/CA-CZB-4091/2020\|EPI_ISL_548634\|2020-0 | ···················· | ·A·················· | ···················· | ···················· | ···················· | ···················· |
| I | 3540 | hCoV-19/USA/CA-CZB-4059/2020\|EPI_ISL_548635\|2020-0 | ···················· | ···················· | ···················· | ···················· | ···················· | ···················· |
| I | 3541 | hCoV-19/USA/CA-CZB-4018/2020\|EPI_ISL_548636\|2020-0 | ···················· | ·A·················· | ···················· | ···················· | ···················· | ···················· |
| I | 3542 | hCoV-19/USA/CA-CZB-4079/2020\|EPI_ISL_548637\|2020-0 | ···················· | ···················· | ···················· | ···················· | ···················· | ···················· |
| I | 3543 | hCoV-19/USA/CA-CZB-4097/2020\|EPI_ISL_548638\|2020-0 | ···················· | ·A·················· | ···················· | ···················· | ···················· | ···················· |
| I | 3544 | hCoV-19/USA/CA-CZB-4015/2020\|EPI_ISL_548639\|2020-0 | ···················· | ·A·················· | ···················· | ···················· | ···················· | ···················· |
| I | 3545 | hCoV-19/USA/CA-CZB-4098/2020\|EPI_ISL_548641\|2020-0 | ···················· | ·A·················· | ···················· | ···················· | ···················· | ···················· |
| I | 3546 | hCoV-19/USA/CA-CZB-4001/2020\|EPI_ISL_548642\|2020-0 | ···················· | ·A·················· | ···················· | ···················· | ···················· | ···················· |
| I | 3547 | hCoV-19/USA/CA-CZB-4008/2020\|EPI_ISL_548643\|2020-0 | ···················· | ·A·················· | ···················· | ···················· | ···················· | ···················· |
| I | 3548 | hCoV-19/USA/CA-CZB-4054/2020\|EPI_ISL_548644\|2020-0 | ···················· | ·A·················· | ···················· | ···················· | ···················· | ···················· |
| I | 3549 | hCoV-19/USA/CA-CZB-4037/2020\|EPI_ISL_548645\|2020-0 | ···················· | ·A·················· | ···················· | ···················· | ···················· | ···················· |
| I | 3550 | hCoV-19/USA/CA-CZB-4095/2020\|EPI_ISL_548646\|2020-0 | ···················· | ·A·················· | ···················· | ···················· | ···················· | ···················· |
| I | 3551 | hCoV-19/USA/CA-CZB-4038/2020\|EPI_ISL_548647\|2020-0 | ···················· | ·A·················· | ···················· | ···················· | ···················· | ···················· |
| I | 3552 | hCoV-19/USA/CA-CZB-4065/2020\|EPI_ISL_548648\|2020-0 | ···················· | ···················· | ···················· | ···················· | ···················· | ···················· |
| I | 3553 | hCoV-19/USA/CA-CZB-4052/2020\|EPI_ISL_548649\|2020-0 | ···················· | ·A·················· | ···················· | ···················· | ···················· | ···················· |
| I | 3554 | hCoV-19/USA/CA-CZB-4031/2020\|EPI_ISL_548650\|2020-0 | ···················· | ·A·················· | ···················· | ···················· | ···················· | ···················· |
| I | 3555 | hCoV-19/USA/CA-CZB-4081/2020\|EPI_ISL_548651\|2020-0 | ···················· | ···················· | ···················· | ···················· | ···················· | ···················· |
| I | 3556 | hCoV-19/USA/CA-CZB-4002/2020\|EPI_ISL_548652\|2020-0 | ···················· | ·A·················· | ···················· | ···················· | ···················· | ···················· |
| I | 3557 | hCoV-19/USA/CA-CZB-4064/2020\|EPI_ISL_548653\|2020-0 | ···················· | ···················· | ···················· | ···················· | ···················· | ···················· |
| I | 3558 | hCoV-19/USA/CA-CZB-4067/2020\|EPI_ISL_548654\|2020-0 | ···················· | ···················· | ···················· | ···················· | ···················· | ···················· |
| I | 3559 | hCoV-19/USA/CA-CZB-4004/2020\|EPI_ISL_548655\|2020-0 | ···················· | ·A·················· | ···················· | ···················· | ···················· | ···················· |
| I | 3560 | hCoV-19/USA/CA-CZB-4006/2020\|EPI_ISL_548656\|2020-0 | ···················· | ·A·················· | ···················· | ···················· | ···················· | ···················· |

I 3561 hCoV-19/USA/CA-CZB-3999/2020|EPI_ISL_548657|2020-0 ···················· ·A·················· ···················· ···················· ···················· ····················
I 3562 hCoV-19/USA/CA-CZB-4020/2020|EPI_ISL_548658|2020-0 ···················· ·A·················· ···················· ···················· ···················· ····················
I 3563 hCoV-19/USA/CA-CZB-4047/2020|EPI_ISL_548659|2020-0 ···················· ···················· ···················· ···················· ···················· ····················
I 3564 hCoV-19/USA/CA-CZB-4077/2020|EPI_ISL_548660|2020-0 ···················· ···················· ···················· ···················· ···················· ····················
I 3565 hCoV-19/USA/CA-CZB-4040/2020|EPI_ISL_548661|2020-0 ···················· ·A·················· ···················· ···················· ···················· ····················
I 3566 hCoV-19/USA/CA-CZB-4062/2020|EPI_ISL_548662|2020-0 ···················· ···················· ···················· ···················· ···················· ····················
I 3567 hCoV-19/USA/CA-CZB-4057/2020|EPI_ISL_548663|2020-0 ···················· ·A·················· ···················· ···················· ···················· ····················
I 3568 hCoV-19/USA/CA-CZB-4030/2020|EPI_ISL_548664|2020-0 ···················· ·A·················· ···················· ···················· ···················· ····················
I 3569 hCoV-19/USA/CA-CZB-4071/2020|EPI_ISL_548665|2020-0 ···················· ···················· ···················· ···················· ···················· ····················
I 3570 hCoV-19/USA/CA-CZB-4099/2020|EPI_ISL_548666|2020-0 ···················· ···················· ···················· ···················· ···K················ ····················
I 3571 hCoV-19/USA/CA-CZB-4075/2020|EPI_ISL_548667|2020-0 ···················· ···················· ···················· ···················· ···················· ····················
I 3572 hCoV-19/USA/CA-CZB-4068/2020|EPI_ISL_548668|2020-0 ···················· ···················· ···················· ···················· ···················· ····················
I 3573 hCoV-19/USA/CA-CZB-4003/2020|EPI_ISL_548669|2020-0 ···················· ·A·················· ···················· ···················· ···················· ····················
I 3574 hCoV-19/USA/CA-CZB-4028/2020|EPI_ISL_548670|2020-0 ···················· ·A·················· ···················· ···················· ···················· ····················
I 3575 hCoV-19/USA/CA-CZB-4056/2020|EPI_ISL_548671|2020-0 ···················· ···················· ···················· ···················· ···················· ····················
I 3576 hCoV-19/USA/CA-CZB-4010/2020|EPI_ISL_548672|2020-0 ···················· ·A·················· ···················· ···················· ···················· ····················
I 3578 hCoV-19/USA/CA-CZB-4069/2020|EPI_ISL_548674|2020-0 ···················· ···················· ···················· ···················· ···················· ····················
I 3579 hCoV-19/USA/CA-CZB-4074/2020|EPI_ISL_548675|2020-0 ···················· ···················· ···················· ···················· ···················· ····················
I 3580 hCoV-19/USA/CA-CZB-4014/2020|EPI_ISL_548676|2020-0 ···················· ·A·················· ···················· ···················· ···················· ····················
I 3581 hCoV-19/USA/CA-CZB-4080/2020|EPI_ISL_548677|2020-0 ···················· ·A·················· ···················· ···················· ···················· ····················
I 3582 hCoV-19/USA/CA-CZB-4023/2020|EPI_ISL_548678|2020-0 ···················· ·A·················· ···················· ···················· ···················· ····················
I 3583 hCoV-19/USA/CA-CZB-4045/2020|EPI_ISL_548679|2020-0 ···················· ·A·················· ···················· ···················· ···················· ····················
I 3584 hCoV-19/Canada/ON-PHL-20-00004/2020|EPI_ISL_548687 ···················· ···················· ···················· ···················· ···················· ····················
I 3585 hCoV-19/USA/TX-HMH-2862/2020|EPI_ISL_543832|2020-0 ···················· ···················· ···················· ···················· ···················· ····················
I 3587 hCoV-19/USA/TX-HMH-2865/2020|EPI_ISL_543835|2020-0 ···················· ···················· ···················· ···················· ···················· ····················
I 3588 hCoV-19/USA/TX-HMH-2866/2020|EPI_ISL_543836|2020-0 ···················· ···················· ···················· ···················· ···················· ····················
I 3591 hCoV-19/USA/TX-HMH-2870/2020|EPI_ISL_543840|2020-0 ···················· ···················· ···················· ···················· ···················· ····················
I 3592 hCoV-19/USA/TX-HMH-2872/2020|EPI_ISL_543841|2020-0 ···················· ···················· ···················· ···················· ···················· ····················
I 3593 hCoV-19/USA/TX-HMH-2873/2020|EPI_ISL_543842|2020-0 ···················· ···················· ···················· ···················· ···················· ····················
I 3594 hCoV-19/USA/TX-HMH-2874/2020|EPI_ISL_543843|2020-0 ···················· ·A·················· ···················· ···················· ···················· ····················
I 3595 hCoV-19/USA/TX-HMH-2875/2020|EPI_ISL_543844|2020-0 ···················· ···················· ···················· ···················· ···················· ····················
I 3596 hCoV-19/USA/TX-HMH-2876/2020|EPI_ISL_543845|2020-0 ···················· ···················· ···················· ···················· ···················· ····················
I 3597 hCoV-19/USA/TX-HMH-2877/2020|EPI_ISL_543846|2020-0 ···················· ···················· ···················· ···················· ···················· ····················
I 3598 hCoV-19/USA/TX-HMH-2878/2020|EPI_ISL_543847|2020-0 ···················· ···················· ···················· ···················· ···················· ····················
I 3599 hCoV-19/USA/TX-HMH-2879/2020|EPI_ISL_543848|2020-0 ···················· ···················· ···················· ···················· ···················· ····················
I 3600 hCoV-19/USA/TX-HMH-2881/2020|EPI_ISL_543849|2020-0 ···················· ···················· ···················· ···················· ···················· ····················
I 3601 hCoV-19/USA/TX-HMH-2882/2020|EPI_ISL_543850|2020-0 ···················· ···················· ···················· ···················· ···················· ····················
I 3602 hCoV-19/USA/TX-HMH-2883/2020|EPI_ISL_543851|2020-0 ···················· ···················· ···················· ···················· ···················· ····················
I 3603 hCoV-19/USA/TX-HMH-2884/2020|EPI_ISL_543852|2020-0 ···················· ···················· ···················· ···················· ···················· ····················
I 3604 hCoV-19/USA/TX-HMH-2885/2020|EPI_ISL_543853|2020-0 ···················· ···················· ···················· ···················· ···················· ····················
I 3605 hCoV-19/USA/TX-HMH-2886/2020|EPI_ISL_543854|2020-0 ···················· ···················· ···················· ···················· ···················· ····················
I 3606 hCoV-19/USA/TX-HMH-2887/2020|EPI_ISL_543855|2020-0 ···················· ···················· ···················· ···················· ···················· ····················
I 3608 hCoV-19/USA/TX-HMH-2889/2020|EPI_ISL_543857|2020-0 ···················· ···················· ···················· ···················· ···················· ····················
I 3609 hCoV-19/USA/TX-HMH-2890/2020|EPI_ISL_543858|2020-0 ···················· ···················· ···················· ···················· ···················· ····················
I 3610 hCoV-19/USA/TX-HMH-2891/2020|EPI_ISL_543859|2020-0 ···················· ···················· ···················· ···················· ···················· ····················
I 3611 hCoV-19/USA/TX-HMH-2892/2020|EPI_ISL_543860|2020-0 ···················· ···················· ···················· ···················· ···················· ····················
I 3612 hCoV-19/USA/TX-HMH-2893/2020|EPI_ISL_543861|2020-0 ···················· ···················· ···················· ···················· ···················· ····················
I 3613 hCoV-19/USA/TX-HMH-2894/2020|EPI_ISL_543862|2020-0 ···················· ···················· ···················· ···················· ···················· ····················
I 3614 hCoV-19/USA/TX-HMH-2920/2020|EPI_ISL_543864|2020-0 ···················· ···················· ···················· ···················· ···················· ····················
I 3615 hCoV-19/USA/TX-HMH-2921/2020|EPI_ISL_543865|2020-0 ···················· ···················· ···················· ···················· ···················· ····················
I 3616 hCoV-19/USA/TX-HMH-2922/2020|EPI_ISL_543866|2020-0 ···················· ···················· ···················· ···················· ···················· ····················
I 3617 hCoV-19/USA/TX-HMH-2926/2020|EPI_ISL_543868|2020-0 ···················· ···················· ···················· ···················· ···················· ····················
I 3618 hCoV-19/USA/TX-HMH-2927/2020|EPI_ISL_543869|2020-0 ···················· ···················· ···················· ···················· ···················· ····················
I 3620 hCoV-19/USA/TX-HMH-2931/2020|EPI_ISL_543872|2020-0 ···················· ···················· ···················· ···················· ···················· ····················
I 3621 hCoV-19/USA/TX-HMH-2934/2020|EPI_ISL_543873|2020-0 ···················· ···················· ···················· ···················· ···················· ······NN·····N······ 1
I 3622 hCoV-19/USA/TX-HMH-2940/2020|EPI_ISL_543876|2020-0 ···················· ···················· ···················· ···················· ···················· ····················
I 3623 hCoV-19/USA/TX-HMH-2943/2020|EPI_ISL_543879|2020-0 ···················· ···················· ···················· ···················· ···················· ····················
I 3624 hCoV-19/USA/TX-HMH-2944/2020|EPI_ISL_543880|2020-0 ···················· ···················· ···················· ···················· ···················· ····················
I 3625 hCoV-19/USA/TX-HMH-2945/2020|EPI_ISL_543881|2020-0 ···················· ···················· ···················· ···················· ···················· ····················
I 3626 hCoV-19/USA/TX-HMH-2946/2020|EPI_ISL_543882|2020-0 ···················· ···················· ···················· ···················· ···················· ····················
I 3627 hCoV-19/USA/TX-HMH-2947/2020|EPI_ISL_543883|2020-0 ···················· ···················· ···················· ···················· ···················· ····················
I 3628 hCoV-19/USA/TX-HMH-2950/2020|EPI_ISL_543886|2020-0 ···················· ···················· ···················· ···················· ···················· ····················
I 3629 hCoV-19/USA/TX-HMH-2954/2020|EPI_ISL_543888|2020-0 ···················· ·············A······ ···················· ···················· ···················· ····················
I 3634 hCoV-19/USA/TX-HMH-2963/2020|EPI_ISL_543897|2020-0 ···················· ···················· ···················· ···················· ···················· ····················
I 3635 hCoV-19/USA/TX-HMH-2964/2020|EPI_ISL_543898|2020-0 ···················· ···················· ···················· ···················· ···················· ····················
I 3636 hCoV-19/USA/TX-HMH-2965/2020|EPI_ISL_543899|2020-0 ···················· ···················· ···················· ···················· ···················· ····················
I 3637 hCoV-19/USA/TX-HMH-3446/2020|EPI_ISL_544200|2020-0 ···················· ···················· ···················· ···················· ···················· NN··················
I 3638 hCoV-19/USA/TX-HMH-3500/2020|EPI_ISL_544203|2020-0 ···················· ···················· ···················· ···················· ···················· ····················
I 3640 hCoV-19/USA/TX-HMH-3505/2020|EPI_ISL_544207|2020-0 ···················· ···················· ···················· ···················· ··············N····· ··N·················
I 3641 hCoV-19/USA/TX-HMH-3506/2020|EPI_ISL_544208|2020-0 ···················· ···················· ···················· ···················· ···················· ····················
I 3645 hCoV-19/USA/TX-HMH-3515/2020|EPI_ISL_544215|2020-0 ···················· ···················· ···················· ···················· ···················· ····················
I 3646 hCoV-19/USA/TX-HMH-3516/2020|EPI_ISL_544216|2020-0 ···················· ···················· ···················· ···················· ···················· ····················
I 3647 hCoV-19/USA/TX-HMH-3519/2020|EPI_ISL_544218|2020-0 ···················· ·A·················· ···················· ···················· ···················· ····················
I 3649 hCoV-19/USA/TX-HMH-3521/2020|EPI_ISL_544220|2020-0 ···················· ···················· ···················· ···················· ···················· NN··················
I 3650 hCoV-19/USA/TX-HMH-3523/2020|EPI_ISL_544221|2020-0 ···················· ···················· ···················· ···················· ···················· ····················
I 3652 hCoV-19/USA/TX-HMH-3525/2020|EPI_ISL_544223|2020-0 ···················· ···················· ···················· ···················· ···················· ····················
I 3653 hCoV-19/USA/TX-HMH-3526/2020|EPI_ISL_544224|2020-0 ···················· ···················· ···················· ···················· ···················· ····················
I 3655 hCoV-19/USA/TX-HMH-3528/2020|EPI_ISL_544226|2020-0 ···················· ···················· ···················· ···················· ···················· ····················
I 3657 hCoV-19/USA/TX-HMH-3532/2020|EPI_ISL_544228|2020-0 ···················· ···················· ···················· ···················· ···················· ····················
I 3664 hCoV-19/USA/TX-HMH-3541/2020|EPI_ISL_544235|2020-0 ···················· ···················· ···················· ···················· ··············N····· NN··················

| | | | | | | | | |
|---|---|---|---|---|---|---|---|---|
| I | 3666 | hCoV-19/USA/TX-HMH-4667/2020\|EPI_ISL_544240\|2020-0 | ···················· | ···················· | ···················· | ···················· | ···················· | ···················· |
| I | 3667 | hCoV-19/USA/TX-HMH-4669/2020\|EPI_ISL_544242\|2020-0 | ···················· | ···················· | ···················· | ···················· | ···················· | ···················· |
| I | 3668 | hCoV-19/USA/TX-HMH-467/2020\|EPI_ISL_544243\|2020-04 | ···················· | ···················· | ···················· | ···················· | ···················· | ···················· |
| I | 3669 | hCoV-19/USA/TX-HMH-4671/2020\|EPI_ISL_544245\|2020-0 | ···················· | ·············A······ | ···················· | ···················· | ···················· | ···················· |
| I | 3670 | hCoV-19/USA/TX-HMH-4672/2020\|EPI_ISL_544246\|2020-0 | ···················· | ···················· | ···················· | ···················· | ···················· | ···················· |
| I | 3671 | hCoV-19/USA/TX-HMH-4673/2020\|EPI_ISL_544247\|2020-0 | ···················· | ···················· | ···················· | ···················· | ···················· | ···················· |
| I | 3674 | hCoV-19/USA/TX-HMH-2752/2020\|EPI_ISL_545528\|2020-0 | ···················· | ···················· | ···················· | ···················· | ···················· | ···················· |
| I | 3676 | hCoV-19/USA/TX-HMH-2756/2020\|EPI_ISL_545531\|2020-0 | ···················· | ···················· | ···················· | ···················· | ···················· | ···················· |
| I | 3677 | hCoV-19/USA/TX-HMH-2757/2020\|EPI_ISL_545532\|2020-0 | ···················· | ···················· | ···················· | ···················· | ···················· | ···················· |
| I | 3678 | hCoV-19/USA/TX-HMH-2758/2020\|EPI_ISL_545533\|2020-0 | ···················· | ···················· | ···················· | ···················· | ···················· | ···················· |
| I | 3680 | hCoV-19/USA/TX-HMH-2763/2020\|EPI_ISL_545537\|2020-0 | ···················· | ···················· | ···················· | ···················· | ···················· | ···················· |
| I | 3681 | hCoV-19/USA/TX-HMH-2764/2020\|EPI_ISL_545538\|2020-0 | ···················· | ···················· | ···················· | ···················· | ···················· | ···················· |
| I | 3682 | hCoV-19/USA/TX-HMH-2765/2020\|EPI_ISL_545539\|2020-0 | ···················· | ···················· | ···················· | ···················· | ···················· | ···················· |
| I | 3683 | hCoV-19/USA/TX-HMH-2767/2020\|EPI_ISL_545540\|2020-0 | ···················· | ···················· | ···················· | ···················· | ···················· | ···················· |
| I | 3684 | hCoV-19/USA/TX-HMH-2768/2020\|EPI_ISL_545541\|2020-0 | ···················· | ···················· | ···················· | ···················· | ···················· | ···················· |
| I | 3685 | hCoV-19/USA/TX-HMH-2769/2020\|EPI_ISL_545542\|2020-0 | ···················· | ······A············· | ···················· | ···················· | ···················· | ···················· |
| I | 3686 | hCoV-19/USA/TX-HMH-2771/2020\|EPI_ISL_545543\|2020-0 | ···················· | ···················· | ···················· | ···················· | ···················· | ···················· |
| I | 3687 | hCoV-19/USA/TX-HMH-2775/2020\|EPI_ISL_545545\|2020-0 | ···················· | ···················· | ···················· | ···················· | ···················· | ···················· |
| I | 3688 | hCoV-19/USA/TX-HMH-2776/2020\|EPI_ISL_545546\|2020-0 | ···················· | ···················· | ···················· | ···················· | ···················· | ···················· |
| I | 3689 | hCoV-19/USA/TX-HMH-2777/2020\|EPI_ISL_545547\|2020-0 | ···················· | ···················· | ···················· | ···················· | ···················· | ···················· |
| I | 3690 | hCoV-19/USA/TX-HMH-2778/2020\|EPI_ISL_545548\|2020-0 | ···················· | ···················· | ···················· | ···················· | ···················· | ···················· |
| I | 3691 | hCoV-19/USA/TX-HMH-2779/2020\|EPI_ISL_545549\|2020-0 | ···················· | ···················· | ···················· | ···················· | ···················· | ···················· |
| I | 3692 | hCoV-19/USA/TX-HMH-2780/2020\|EPI_ISL_545550\|2020-0 | ···················· | ···················· | ···················· | ···················· | ···················· | ···················· |
| I | 3693 | hCoV-19/USA/TX-HMH-2787/2020\|EPI_ISL_545552\|2020-0 | ···················· | ······A············· | ···················· | ···················· | ···················· | ···················· |
| I | 3694 | hCoV-19/USA/TX-HMH-2788/2020\|EPI_ISL_545553\|2020-0 | ···················· | ···················· | ···················· | ···················· | ···················· | ···················· |
| I | 3695 | hCoV-19/USA/TX-HMH-2790/2020\|EPI_ISL_545555\|2020-0 | ···················· | ·A·················· | ···················· | ···················· | ···················· | ···················· |
| I | 3696 | hCoV-19/USA/TX-HMH-2791/2020\|EPI_ISL_545556\|2020-0 | ···················· | ······A············· | ···················· | ···················· | ···················· | ···················· |
| I | 3697 | hCoV-19/USA/TX-HMH-2792/2020\|EPI_ISL_545557\|2020-0 | ···················· | ···················· | ···················· | ···················· | ···················· | ···················· |
| I | 3698 | hCoV-19/USA/TX-HMH-2793/2020\|EPI_ISL_545558\|2020-0 | ···················· | ···················· | ···················· | ···················· | ···················· | ···················· |
| I | 3699 | hCoV-19/USA/TX-HMH-2795/2020\|EPI_ISL_545559\|2020-0 | ···················· | ······A············· | ···················· | ···················· | ···················· | ···················· |
| I | 3700 | hCoV-19/USA/TX-HMH-2797/2020\|EPI_ISL_545560\|2020-0 | ···················· | ···················· | ···················· | ···················· | ···················· | ···················· |
| I | 3701 | hCoV-19/USA/TX-HMH-2798/2020\|EPI_ISL_545561\|2020-0 | ···················· | ···················· | ···················· | ···················· | ···················· | ···················· |
| I | 3702 | hCoV-19/USA/TX-HMH-2799/2020\|EPI_ISL_545562\|2020-0 | ···················· | ···················· | ···················· | ···················· | ···················· | ···················· |
| I | 3703 | hCoV-19/USA/TX-HMH-2800/2020\|EPI_ISL_545563\|2020-0 | ···················· | ······A············· | ···················· | ···················· | ···················· | ···················· |
| I | 3704 | hCoV-19/USA/TX-HMH-2801/2020\|EPI_ISL_545564\|2020-0 | ···················· | ···················· | ···················· | ···················· | ···················· | ···················· |
| I | 3705 | hCoV-19/USA/TX-HMH-2802/2020\|EPI_ISL_545565\|2020-0 | ···················· | ···················· | ···················· | ···················· | ···················· | ···················· |
| I | 3706 | hCoV-19/USA/TX-HMH-2804/2020\|EPI_ISL_545566\|2020-0 | ···················· | ···················· | ···················· | ···················· | ···················· | ···················· |
| I | 3707 | hCoV-19/USA/TX-HMH-2805/2020\|EPI_ISL_545567\|2020-0 | ···················· | ···················· | ···················· | ···················· | ···················· | ···················· |
| I | 3709 | hCoV-19/USA/TX-HMH-2807/2020\|EPI_ISL_545569\|2020-0 | ···················· | ···················· | ···················· | ···················· | ···················· | ···················· |
| I | 3710 | hCoV-19/USA/TX-HMH-2808/2020\|EPI_ISL_545570\|2020-0 | ···················· | ···················· | ···················· | ···················· | ···················· | ···················· |
| I | 3711 | hCoV-19/USA/TX-HMH-2811/2020\|EPI_ISL_545573\|2020-0 | ···················· | ·············A······ | ···················· | ···················· | ···················· | ···················· |
| I | 3712 | hCoV-19/USA/TX-HMH-2812/2020\|EPI_ISL_545574\|2020-0 | ···················· | ···················· | ···················· | ···················· | ···················· | ···················· |
| I | 3713 | hCoV-19/USA/TX-HMH-7719/2020\|EPI_ISL_545600\|2020-0 | ···················· | ···················· | ···················· | ···················· | ···················· | ···················· |
| I | 3714 | hCoV-19/USA/TX-HMH-772/2020\|EPI_ISL_545601\|2020-04 | ···················· | ···················· | ···················· | ···················· | ···················· | ···················· |
| I | 3715 | hCoV-19/USA/TX-HMH-7723/2020\|EPI_ISL_545602\|2020-0 | ···················· | ···················· | ···················· | ···················· | ···················· | ···················· |
| I | 3716 | hCoV-19/USA/TX-HMH-7732/2020\|EPI_ISL_545606\|2020-0 | ···················· | ···················· | ···················· | ···················· | ···················· | ···················· |
| I | 3718 | hCoV-19/USA/TX-HMH-7736/2020\|EPI_ISL_545608\|2020-0 | ···················· | ···················· | ···················· | ···················· | ···················· | ···················· |
| I | 3719 | hCoV-19/USA/TX-HMH-7739/2020\|EPI_ISL_545609\|2020-0 | ···················· | ···················· | ···················· | ···················· | ···················· | ···················· |
| I | 3720 | hCoV-19/USA/TX-HMH-774/2020\|EPI_ISL_545610\|2020-04 | ···················· | ···················· | ···················· | ···················· | ···················· | ···················· |
| I | 3721 | hCoV-19/USA/TX-HMH-7740/2020\|EPI_ISL_545611\|2020-0 | ···················· | ··········A········· | ···················· | ···················· | ···················· | ···················· |
| I | 3723 | hCoV-19/USA/TX-HMH-6487/2020\|EPI_ISL_546031\|2020-0 | ···················· | ···················· | ···················· | ···················· | ··············N····· | ···················· |
| I | 3725 | hCoV-19/USA/TX-HMH-6489/2020\|EPI_ISL_546033\|2020-0 | ···················· | ······A············· | ···················· | ···················· | ···················· | ···················· |
| I | 3727 | hCoV-19/USA/TX-HMH-6493/2020\|EPI_ISL_546036\|2020-0 | ···················· | ···················· | ···················· | ···················· | ···················· | ···················· |
| I | 3728 | hCoV-19/USA/TX-HMH-6496/2020\|EPI_ISL_546037\|2020-0 | ···················· | ·A·················· | ···················· | ···················· | ···················· | ···················· |
| I | 3729 | hCoV-19/USA/TX-HMH-6497/2020\|EPI_ISL_546038\|2020-0 | ···················· | ···················· | ···················· | ···················· | ···················· | ···················· |
| I | 3730 | hCoV-19/USA/TX-HMH-6499/2020\|EPI_ISL_546039\|2020-0 | ···················· | ···················· | ···················· | ···················· | ···················· | ···················· |
| I | 3731 | hCoV-19/USA/TX-HMH-6500/2020\|EPI_ISL_546040\|2020-0 | ···················· | ···················· | ···················· | ···················· | ···················· | ···················· |
| I | 3732 | hCoV-19/USA/TX-HMH-6502/2020\|EPI_ISL_546041\|2020-0 | ···················· | ···················· | ···················· | ···················· | ···················· | ···················· |
| I | 3733 | hCoV-19/USA/TX-HMH-6503/2020\|EPI_ISL_546042\|2020-0 | ···················· | ···················· | ···················· | ···················· | ···················· | ···················· |
| I | 3734 | hCoV-19/USA/TX-HMH-6504/2020\|EPI_ISL_546043\|2020-0 | ···················· | ···················· | ···················· | ···················· | ···················· | ···················· |
| I | 3735 | hCoV-19/USA/TX-HMH-6505/2020\|EPI_ISL_546044\|2020-0 | ···················· | ···················· | ···················· | ···················· | ···················· | ···················· |
| I | 3736 | hCoV-19/USA/TX-HMH-6506/2020\|EPI_ISL_546045\|2020-0 | ···················· | ···················· | ···················· | ···················· | ···················· | ·······NN··········· |
| I | 3737 | hCoV-19/USA/TX-HMH-6507/2020\|EPI_ISL_546046\|2020-0 | ···················· | ·A·················· | ···················· | ···················· | ···················· | ···················· |
| I | 3738 | hCoV-19/USA/TX-HMH-6510/2020\|EPI_ISL_546047\|2020-0 | ···················· | ···················· | ···················· | ···················· | ···················· | ···················· |
| I | 3739 | hCoV-19/USA/TX-HMH-6511/2020\|EPI_ISL_546048\|2020-0 | ···················· | ···················· | ···················· | ···················· | ···················· | ···N················ |
| I | 3740 | hCoV-19/USA/TX-HMH-6513/2020\|EPI_ISL_546049\|2020-0 | ···················· | ···················· | ···················· | ···················· | ···················· | ···················· |
| I | 3741 | hCoV-19/USA/TX-HMH-6516/2020\|EPI_ISL_546051\|2020-0 | ···················· | ···················· | ···················· | ···················· | ···················· | ···················· |
| I | 3742 | hCoV-19/USA/TX-HMH-6517/2020\|EPI_ISL_546052\|2020-0 | ···················· | ···················· | ···················· | ···················· | ···················· | ···················· |
| I | 3743 | hCoV-19/USA/TX-HMH-6519/2020\|EPI_ISL_546053\|2020-0 | ···················· | ···················· | ···················· | ···················· | ···················· | ···················· |
| I | 3744 | hCoV-19/USA/TX-HMH-6522/2020\|EPI_ISL_546055\|2020-0 | ···················· | ···················· | ···················· | ···················· | ···················· | ···················· |
| I | 3745 | hCoV-19/USA/TX-HMH-6523/2020\|EPI_ISL_546056\|2020-0 | ···················· | ···················· | ···················· | ···················· | ···················· | ···················· |
| I | 3748 | hCoV-19/USA/TX-HMH-7570/2020\|EPI_ISL_546861\|2020-0 | ···················· | ···················· | ···················· | ···················· | ···················· | ·······N············ |
| I | 3749 | hCoV-19/USA/TX-HMH-7571/2020\|EPI_ISL_546862\|2020-0 | ···················· | ···················· | ···················· | ···················· | ···················· | ···················· |
| I | 3751 | hCoV-19/USA/TX-HMH-7575/2020\|EPI_ISL_546864\|2020-0 | ···················· | ···················· | ···················· | ···················· | ···················· | ···················· |
| I | 3753 | hCoV-19/USA/TX-HMH-758/2020\|EPI_ISL_546868\|2020-04 | ···················· | ···················· | ···················· | ···················· | ···················· | ···················· |
| I | 3754 | hCoV-19/USA/TX-HMH-7580/2020\|EPI_ISL_546869\|2020-0 | ···················· | ···················· | ···················· | ···················· | ···················· | ···················· |
| I | 3756 | hCoV-19/USA/TX-HMH-7583/2020\|EPI_ISL_546871\|2020-0 | ···················· | ···················· | ···················· | ···················· | ···················· | ···················· |
| I | 3758 | hCoV-19/USA/TX-HMH-7594/2020\|EPI_ISL_546879\|2020-0 | ···················· | ···················· | ···················· | ···················· | ···················· | ···················· |
| I | 3759 | hCoV-19/USA/TX-HMH-7597/2020\|EPI_ISL_546880\|2020-0 | ···················· | ···················· | ···················· | ···················· | ···················· | ···················· |
| I | 3762 | hCoV-19/USA/TX-HMH-7602/2020\|EPI_ISL_546884\|2020-0 | ···················· | ···················· | ···················· | ···················· | ···················· | ···N················ |

```
I 3763 hCoV-19/USA/TX-HMH-7603/2020|EPI_ISL_546885|2020-0 ···················· ···················· ····················· ···················· ···················· ····················
I 3767 hCoV-19/USA/TX-HMH-7609/2020|EPI_ISL_546889|2020-0 ···················· ···················· ····················· ···················· ···················· ····················
I 3769 hCoV-19/USA/TX-HMH-7619/2020|EPI_ISL_546892|2020-0 ···················· ·············A······ ····················· ···················· ···················· ····················
I 3770 hCoV-19/USA/TX-HMH-7620/2020|EPI_ISL_546893|2020-0 ···················· ···················· ····················· ···················· ···················· ····················
I 3772 hCoV-19/USA/TX-HMH-7625/2020|EPI_ISL_546895|2020-0 ···················· ···················· ····················· ···················· ···················· ····················
I 3773 hCoV-19/USA/TX-HMH-7626/2020|EPI_ISL_546896|2020-0 ···················· ···················· ····················· ···················· ··············N····· N···················
I 3774 hCoV-19/USA/TX-HMH-7627/2020|EPI_ISL_546897|2020-0 ···················· ·A·················· ····················· ···················· ···················· ····················
I 3777 hCoV-19/USA/TX-HMH-7630/2020|EPI_ISL_546900|2020-0 ···················· ···················· ····················· ···················· ···················· ····················
I 3778 hCoV-19/USA/TX-HMH-7633/2020|EPI_ISL_546902|2020-0 ···················· ···················· ····················· ···················· ···················· ····················
I 3779 hCoV-19/USA/TX-HMH-7635/2020|EPI_ISL_546904|2020-0 ···················· ···················· ····················· ···················· ···················· ····················
I 3780 hCoV-19/USA/TX-HMH-7636/2020|EPI_ISL_546905|2020-0 ···················· ···················· ····················· ···················· ···················· ····················
I 3781 hCoV-19/USA/TX-HMH-7639/2020|EPI_ISL_546906|2020-0 ···················· ···················· ····················· ···················· ··············N····· ····················
I 3782 hCoV-19/USA/TX-HMH-7641/2020|EPI_ISL_546907|2020-0 ···················· ···················· ····················· ···················· ···················· ····················
I 3783 hCoV-19/USA/TX-HMH-7642/2020|EPI_ISL_546908|2020-0 ···················· ·············A······ ····················· ···················· ···················· ····················
I 3784 hCoV-19/USA/TX-HMH-7643/2020|EPI_ISL_546909|2020-0 ···················· ···················· ····················· ···················· ···················· ····················
I 3786 hCoV-19/USA/TX-HMH-7646/2020|EPI_ISL_546911|2020-0 ···················· ···················· ····················· ···················· ···················· ····················
I 3788 hCoV-19/USA/TX-HMH-765/2020|EPI_ISL_546913|2020-04 ···················· ···················· ····················· ···················· ···················· ····················
I 3789 hCoV-19/USA/TX-HMH-7650/2020|EPI_ISL_546914|2020-0 ···················· ··········A········· ····················· ···················· ···················· ····················
I 3792 hCoV-19/USA/TX-HMH-7658/2020|EPI_ISL_546920|2020-0 ···················· ·············A······ ····················· ···················· ···················· ····················
I 3793 hCoV-19/USA/TX-HMH-7659/2020|EPI_ISL_546921|2020-0 ···················· ···················· ····················· ···················· ··············N····· ·······NN···········
I 3798 hCoV-19/USA/TX-HMH-7676/2020|EPI_ISL_546932|2020-0 ···················· ···················· ····················· ···················· ···················· ····················
I 3800 hCoV-19/Czech Republic/NRL-8118-1/2020|EPI_ISL_546 ···················· ···················· ····················· ···················· ···················· ····················
I 3801 hCoV-19/Czech Republic/NRL-8118-2/2020|EPI_ISL_546 ···················· ···················· ····················· ···················· ···················· ····················
I 3804 hCoV-19/USA/TX-HMH-568/2020|EPI_ISL_546940|2020-04 ···················· ···················· ····················· ···················· ···················· ····················
I 3806 hCoV-19/USA/TX-HMH-6279/2020|EPI_ISL_547360|2020-0 ···················· ···················· ····················· ···················· ···················· ····················
I 3808 hCoV-19/USA/TX-HMH-6284/2020|EPI_ISL_547363|2020-0 ···················· ···················· ····················· ···················· ···················· ····················
I 3809 hCoV-19/USA/TX-HMH-6286/2020|EPI_ISL_547365|2020-0 ···················· ······A············· ····················· ···················· ···················· ····················
I 3813 hCoV-19/USA/TX-HMH-6298/2020|EPI_ISL_547372|2020-0 ···················· ···················· ····················· ···················· ···················· ····················
I 3814 hCoV-19/USA/TX-HMH-6299/2020|EPI_ISL_547373|2020-0 ···················· ···················· ····················· ···················· ···················· ····················
I 3816 hCoV-19/USA/TX-HMH-6301/2020|EPI_ISL_547375|2020-0 ···················· ···················· ····················· ···················· ···················· ····················
I 3817 hCoV-19/USA/TX-HMH-6302/2020|EPI_ISL_547376|2020-0 ···················· ···················· ····················· ···················· ···················· ····················
I 3818 hCoV-19/USA/TX-HMH-6304/2020|EPI_ISL_547378|2020-0 ···················· ···················· ····················· ···················· ···················· ····················
I 3819 hCoV-19/USA/TX-HMH-6305/2020|EPI_ISL_547379|2020-0 ···················· ···················· ····················· ···················· ···················· NN··················
I 3820 hCoV-19/USA/TX-HMH-6307/2020|EPI_ISL_547380|2020-0 ···················· ···················· ····················· ···················· ···················· ····················
I 3821 hCoV-19/USA/TX-HMH-6308/2020|EPI_ISL_547381|2020-0 ···················· ···················· ····················· ···················· ···················· ····················
I 3823 hCoV-19/USA/TX-HMH-6312/2020|EPI_ISL_547383|2020-0 ···················· ···················· ····················· ···················· ···················· ····················
I 3824 hCoV-19/USA/TX-HMH-6315/2020|EPI_ISL_547384|2020-0 ···················· ···················· ····················· ···················· ···················· ····················
I 3825 hCoV-19/USA/TX-HMH-6318/2020|EPI_ISL_547387|2020-0 ···················· ···················· ····················· ···················· ··············N····· ·······NN···········
I 3826 hCoV-19/USA/TX-HMH-6319/2020|EPI_ISL_547388|2020-0 ···················· ···················· ····················· ···················· ···················· ····················
I 3827 hCoV-19/USA/CA-CZB-4094/2020|EPI_ISL_548601|2020-0 ···················· ·A·················· ····················· ···················· ···················· ····················
I 3828 hCoV-19/USA/CA-CZB-4049/2020|EPI_ISL_548602|2020-0 ···················· ···················· ····················· ···················· ···················· ····················
I 3829 hCoV-19/USA/CA-CZB-4055/2020|EPI_ISL_548603|2020-0 ···················· ···················· ····················· ···················· ···················· ····················
I 3830 hCoV-19/USA/CA-CZB-3998/2020|EPI_ISL_548604|2020-0 ···················· ·A·················· ····················· ···················· ···················· ····················
I 3831 hCoV-19/Canada/ON-PHL-20-00007/2020|EPI_ISL_548690 ···················· ···················· ····················· ···················· ···················· ····················
I 3832 hCoV-19/Canada/ON-PHL-20-00008/2020|EPI_ISL_548691 ···················· ···················· ····················· ···················· ···················· ····················
I 3833 hCoV-19/Canada/ON-PHL-20-00009/2020|EPI_ISL_548692 ···················· ···················· ····················· ···················· ···················· ····················
I 3834 hCoV-19/Canada/ON-PHL-20-00010/2020|EPI_ISL_548693 ···················· ···················· ····················· ···················· ···················· ····················
I 3835 hCoV-19/Canada/ON-PHL-20-00011/2020|EPI_ISL_548694 ···················· ···················· ····················· ···················· ···················· ····················
I 3836 hCoV-19/Canada/ON-PHL-20-00012/2020|EPI_ISL_548695 ···················· ···················· ····················· ···················· ···················· ····················
I 3837 hCoV-19/Canada/ON-PHL-20-00014/2020|EPI_ISL_548696 ···················· ···················· ····················· ···················· ···················· ····················
I 3838 hCoV-19/Canada/ON-PHL-20-00015/2020|EPI_ISL_548697 ···················· ···················· ····················· ···················· ···················· ····················
I 3839 hCoV-19/Canada/ON-PHL-20-00016/2020|EPI_ISL_548698 ···················· ···················· ····················· ···················· ···················· ····················
I 3840 hCoV-19/Canada/ON-PHL-20-00018/2020|EPI_ISL_548699 ···················· ···················· ····················· ···················· ···················· ····················
I 3841 hCoV-19/USA/TX-HMH-2827/2020|EPI_ISL_543801|2020-0 ···················· ···················· ····················· ···················· ···················· ····················
I 3842 hCoV-19/USA/TX-HMH-2828/2020|EPI_ISL_543802|2020-0 ···················· ···················· ····················· ···················· ···················· ····················
I 3843 hCoV-19/USA/TX-HMH-2829/2020|EPI_ISL_543803|2020-0 ···················· ···················· ····················· ···················· ···················· ····················
I 3844 hCoV-19/USA/TX-HMH-2830/2020|EPI_ISL_543804|2020-0 ···················· ···················· ····················· ···················· ···················· ····················
I 3845 hCoV-19/USA/TX-HMH-2832/2020|EPI_ISL_543805|2020-0 ···················· ······A············· ····················· ···················· ···················· ····················
I 3846 hCoV-19/USA/TX-HMH-2833/2020|EPI_ISL_543806|2020-0 ···················· ···················· ····················· ···················· ···················· ····················
I 3847 hCoV-19/USA/TX-HMH-2834/2020|EPI_ISL_543807|2020-0 ···················· ···················· ····················· ···················· ···················· ····················
I 3850 hCoV-19/USA/TX-HMH-2839/2020|EPI_ISL_543810|2020-0 ···················· ···················· ····················· ···················· ···················· ····················
I 3851 hCoV-19/USA/TX-HMH-2840/2020|EPI_ISL_543811|2020-0 ···················· ···················· ····················· ···················· ···················· ····················
I 3852 hCoV-19/USA/TX-HMH-2841/2020|EPI_ISL_543812|2020-0 ···················· ·············A······ ····················· ···················· ···················· ····················
I 3853 hCoV-19/USA/TX-HMH-2842/2020|EPI_ISL_543813|2020-0 ···················· ···················· ····················· ···················· ···················· ····················
I 3854 hCoV-19/USA/TX-HMH-2843/2020|EPI_ISL_543814|2020-0 ···················· ···················· ····················· ···················· ···················· ····················
I 3855 hCoV-19/USA/TX-HMH-2844/2020|EPI_ISL_543815|2020-0 ···················· ···················· ····················· ···················· ···················· ····················
I 3856 hCoV-19/USA/TX-HMH-2845/2020|EPI_ISL_543816|2020-0 ···················· ···················· ····················· ···················· ··············NN···· ····················
I 3857 hCoV-19/USA/TX-HMH-2846/2020|EPI_ISL_543817|2020-0 ···················· ···················· ····················· ···················· ··············N····· ····················
I 3858 hCoV-19/USA/TX-HMH-2847/2020|EPI_ISL_543818|2020-0 ···················· ···················· ····················· ···················· ···················· ····················
I 3862 hCoV-19/USA/TX-HMH-2852/2020|EPI_ISL_543823|2020-0 ···················· ······A············· ····················· ···················· N·············NN···· NN·····N············ 1
I 3864 hCoV-19/USA/TX-HMH-2857/2020|EPI_ISL_543827|2020-0 ···················· ···················· ····················· ···················· ···················· ····················
I 3866 hCoV-19/USA/TX-HMH-2859/2020|EPI_ISL_543829|2020-0 ···················· ······A············· ····················· ···················· ···················· ····················
I 3868 hCoV-19/USA/TX-HMH-6525/2020|EPI_ISL_546058|2020-0 ···················· ···················· ····················· ···················· ···················· ····················
I 3869 hCoV-19/USA/TX-HMH-6526/2020|EPI_ISL_546059|2020-0 ···················· ·············A······ ····················· ···················· ···················· ····················
I 3870 hCoV-19/USA/TX-HMH-6529/2020|EPI_ISL_546060|2020-0 ···················· ···················· ····················· ···················· ···················· ····················
I 3871 hCoV-19/USA/TX-HMH-6530/2020|EPI_ISL_546061|2020-0 ···················· ···················· ····················· ···················· ···················· ····················
I 3872 hCoV-19/USA/TX-HMH-6531/2020|EPI_ISL_546062|2020-0 ···················· ···················· ····················· ···················· ···················· ····················
I 3873 hCoV-19/USA/TX-HMH-6535/2020|EPI_ISL_546064|2020-0 ···················· ···················· ····················· ···················· ··············N····· NN·N················ 1
I 3874 hCoV-19/USA/TX-HMH-6538/2020|EPI_ISL_546065|2020-0 ···················· ···················· ····················· ···················· ···················· ····················
I 3875 hCoV-19/USA/TX-HMH-6540/2020|EPI_ISL_546066|2020-0 ···················· ···················· ····················· ···················· ···················· ····················
```

| | | | | | | | | |
|---|---|---|---|---|---|---|---|---|
| I | 3876 | hCoV-19/USA/TX-HMH-6541/2020\|EPI_ISL_546067\|2020-0 | ···················· | ···················· | ····················· | ···················· | ···················· | ···················· |
| I | 3878 | hCoV-19/USA/TX-HMH-6543/2020\|EPI_ISL_546069\|2020-0 | ···················· | ···················· | ····················· | ···················· | ···················· | ···················· |
| I | 3879 | hCoV-19/USA/TX-HMH-6544/2020\|EPI_ISL_546070\|2020-0 | ···················· | ···················· | ····················· | ···················· | ···················· | ···················· |
| I | 3880 | hCoV-19/USA/TX-HMH-6545/2020\|EPI_ISL_546071\|2020-0 | ···················· | ···················· | ····················· | ···················· | ···················· | ···················· |
| I | 3881 | hCoV-19/USA/TX-HMH-6546/2020\|EPI_ISL_546072\|2020-0 | ···················· | ·············A······ | ····················· | ···················· | ···················· | ···················· |
| I | 3882 | hCoV-19/USA/TX-HMH-6547/2020\|EPI_ISL_546073\|2020-0 | ···················· | ···················· | ····················· | ···················· | ···················· | NN·················· |
| I | 3886 | hCoV-19/USA/TX-HMH-6555/2020\|EPI_ISL_546080\|2020-0 | ···················· | ···················· | ····················· | ···················· | ···················· | ···················· |
| I | 3887 | hCoV-19/USA/TX-HMH-6556/2020\|EPI_ISL_546081\|2020-0 | ···················· | ···················· | ····················· | ···················· | ···················· | ···················· |
| I | 3888 | hCoV-19/USA/TX-HMH-6557/2020\|EPI_ISL_546082\|2020-0 | ···················· | ···················· | ····················· | ···················· | ···················· | ···················· |
| I | 3889 | hCoV-19/USA/TX-HMH-6558/2020\|EPI_ISL_546083\|2020-0 | ···················· | ···················· | ····················· | ···················· | ···················· | ···················· |
| I | 3890 | hCoV-19/USA/TX-HMH-6559/2020\|EPI_ISL_546084\|2020-0 | ···················· | ···················· | ····················· | ···················· | ···················· | ···················· |
| I | 3891 | hCoV-19/USA/TX-HMH-6564/2020\|EPI_ISL_546088\|2020-0 | ···················· | ···················· | ····················· | ···················· | ···················· | ···················· |
| I | 3892 | hCoV-19/USA/TX-HMH-6566/2020\|EPI_ISL_546090\|2020-0 | ···················· | ···················· | ····················· | ···················· | ···················· | ···················· |
| I | 3893 | hCoV-19/USA/TX-HMH-6568/2020\|EPI_ISL_546091\|2020-0 | ···················· | ···················· | ····················· | ···················· | ···················· | ···················· |
| I | 3894 | hCoV-19/USA/TX-HMH-6570/2020\|EPI_ISL_546093\|2020-0 | ···················· | ···················· | ····················· | ···················· | ···················· | ···················· |
| I | 3895 | hCoV-19/USA/TX-HMH-6572/2020\|EPI_ISL_546095\|2020-0 | ···················· | ···················· | ····················· | ···················· | ···················· | N··················· |
| I | 3896 | hCoV-19/USA/TX-HMH-6573/2020\|EPI_ISL_546096\|2020-0 | ···················· | ···················· | ····················· | ···················· | ···················· | ···················· |
| I | 3897 | hCoV-19/USA/TX-HMH-6575/2020\|EPI_ISL_546097\|2020-0 | ···················· | ···················· | ····················· | ···················· | ···················· | ···················· |
| I | 3898 | hCoV-19/USA/TX-HMH-6577/2020\|EPI_ISL_546098\|2020-0 | ···················· | ···················· | ····················· | ···················· | ···················· | ···················· |
| I | 3899 | hCoV-19/USA/TX-HMH-6578/2020\|EPI_ISL_546099\|2020-0 | ···················· | ···················· | ····················· | ···················· | ···················· | ···················· |
| I | 3900 | hCoV-19/USA/TX-HMH-6322/2020\|EPI_ISL_547390\|2020-0 | ···················· | ···················· | ····················· | ···················· | ···················· | ···················· |
| I | 3901 | hCoV-19/USA/TX-HMH-6330/2020\|EPI_ISL_547394\|2020-0 | ···················· | ···················· | ····················· | ···················· | ···················· | ···················· |
| I | 3902 | hCoV-19/USA/TX-HMH-6331/2020\|EPI_ISL_547395\|2020-0 | ···················· | ···················· | ····················· | ···················· | ···················· | ···················· |
| I | 3904 | hCoV-19/USA/TX-HMH-6333/2020\|EPI_ISL_547397\|2020-0 | ···················· | ···················· | ····················· | ···················· | ···················· | ···················· |
| I | 3905 | hCoV-19/USA/TX-HMH-6335/2020\|EPI_ISL_547399\|2020-0 | ···················· | ···················· | ····················· | ···················· | ···················· | ···················· |
| J | 1 | hCoV-19/Sweden/RV-FOI-5/2020\|EPI_ISL_549024\|2020-0 | ···················· | ···················· | ····················· | ···················· | ···················· | ···················· |
| J | 2 | hCoV-19/Norway/2952/2020\|EPI_ISL_549027\|2020-07-31 | ···················· | ···················· | ····················· | ···················· | ···················· | ···················· |
| J | 3 | hCoV-19/Norway/3195/2020\|EPI_ISL_549028\|2020-08-26 | ···················· | ···················· | ····················· | ···················· | ···················· | ···················· |
| J | 4 | hCoV-19/Norway/3197/2020\|EPI_ISL_549029\|2020-08-19 | ···················· | ···················· | ····················· | ···················· | ···················· | ···················· |
| J | 5 | hCoV-19/England/MILK-9A6944/2020\|EPI_ISL_550002\|20 | ···················· | ···················· | ····················· | ···················· | ···················· | ···················· |
| J | 6 | hCoV-19/Wales/MILK-9A572D/2020\|EPI_ISL_550003\|2020 | ···················· | ···················· | ····················· | ···················· | ···················· | ···················· |
| J | 7 | hCoV-19/England/MILK-9A8511/2020\|EPI_ISL_550004\|20 | ···················· | ···················· | ····················· | ···················· | ···················· | ···················· |
| J | 8 | hCoV-19/England/MILK-9A5031/2020\|EPI_ISL_550005\|20 | ···················· | ···················· | ····················· | ···················· | ···················· | ···················· |
| J | 9 | hCoV-19/England/MILK-9A6E45/2020\|EPI_ISL_550006\|20 | ···················· | ···················· | ····················· | ···················· | ···················· | ···················· |
| J | 10 | hCoV-19/England/MILK-9A85D5/2020\|EPI_ISL_550007\|20 | ···················· | ···················· | ····················· | ···················· | ···················· | ···················· |
| J | 11 | hCoV-19/England/MILK-9A5356/2020\|EPI_ISL_550008\|20 | ···················· | ···················· | ····················· | ···················· | ···················· | ···················· |
| J | 12 | hCoV-19/England/MILK-9A84E7/2020\|EPI_ISL_550009\|20 | ···················· | ···················· | ····················· | ···················· | ···················· | ···················· |
| J | 13 | hCoV-19/England/MILK-9A5426/2020\|EPI_ISL_550011\|20 | ···················· | ···················· | ····················· | ···················· | ···················· | ···················· |
| J | 14 | hCoV-19/England/MILK-9A5480/2020\|EPI_ISL_550012\|20 | ···················· | ·A·················· | ····················· | ···················· | ···················· | ···················· |
| J | 15 | hCoV-19/England/MILK-9A66E3/2020\|EPI_ISL_550020\|20 | ···················· | ···················· | ····················· | ···················· | ···················· | ···················· |
| J | 16 | hCoV-19/England/ALDP-952349/2020\|EPI_ISL_551333\|20 | ···················· | ·A·················· | ····················· | ···················· | ···················· | ···················· |
| J | 17 | hCoV-19/England/ALDP-9520F7/2020\|EPI_ISL_551334\|20 | ···················· | ···················· | ····················· | ···················· | ···················· | ···················· |
| J | 18 | hCoV-19/England/ALDP-951B24/2020\|EPI_ISL_551335\|20 | ···················· | ·A·················· | ····················· | ···················· | ···················· | ···················· |
| J | 19 | hCoV-19/England/ALDP-951E2B/2020\|EPI_ISL_551338\|20 | ···················· | ···················· | ····················· | ···················· | ···················· | ···················· |
| J | 20 | hCoV-19/England/ALDP-951DC4/2020\|EPI_ISL_551339\|20 | ···················· | ···················· | ····················· | ···················· | ···················· | ···················· |
| J | 21 | hCoV-19/England/ALDP-951DF1/2020\|EPI_ISL_551340\|20 | ···················· | ···················· | ····················· | ···················· | ···················· | ···················· |
| J | 22 | hCoV-19/England/ALDP-951F0A/2020\|EPI_ISL_551341\|20 | ···················· | ···················· | ····················· | ···················· | ···················· | ···················· |
| J | 23 | hCoV-19/England/ALDP-951F91/2020\|EPI_ISL_551342\|20 | ···················· | ···················· | ····················· | ···················· | ···················· | ···················· |
| J | 24 | hCoV-19/England/ALDP-952121/2020\|EPI_ISL_551343\|20 | ···················· | ···················· | ····················· | ···················· | ···················· | ···················· |
| J | 25 | hCoV-19/England/MILK-975BB1/2020\|EPI_ISL_552664\|20 | ···················· | ···················· | ····················· | ···················· | ···················· | ···················· |
| J | 26 | hCoV-19/England/ALDP-98295D/2020\|EPI_ISL_552665\|20 | ···················· | ···················· | ····················· | ···················· | ···················· | ···················· |
| J | 27 | hCoV-19/England/MILK-975AE1/2020\|EPI_ISL_552667\|20 | ···················· | ···················· | ····················· | ···················· | ···················· | ···················· |
| J | 28 | hCoV-19/Wales/MILK-975647/2020\|EPI_ISL_552668\|2020 | ···················· | ···················· | ····················· | ···················· | ···················· | ···················· |
| J | 29 | hCoV-19/England/ALDP-982B84/2020\|EPI_ISL_552669\|20 | ···················· | ···················· | ····················· | ···················· | ···················· | ···················· |
| J | 30 | hCoV-19/England/ALDP-982DBB/2020\|EPI_ISL_552671\|20 | ···················· | ···················· | ····················· | ···················· | ···················· | ···················· |
| J | 31 | hCoV-19/England/MILK-974E5F/2020\|EPI_ISL_552672\|20 | ···················· | ···················· | ····················· | ···················· | ···················· | ···················· |
| J | 32 | hCoV-19/England/MILK-9756A1/2020\|EPI_ISL_552673\|20 | ···················· | ·A·················· | ····················· | ···················· | ···················· | ···················· |
| J | 33 | hCoV-19/England/MILK-975920/2020\|EPI_ISL_552674\|20 | ···················· | ···················· | ····················· | ···················· | ···················· | ···················· |
| J | 34 | hCoV-19/England/MILK-974F89/2020\|EPI_ISL_552683\|20 | ···················· | ···················· | ····················· | ···················· | ···················· | ···················· |
| J | 35 | hCoV-19/England/ALDP-955AF6/2020\|EPI_ISL_553900\|20 | ···················· | ···················· | ····················· | ···················· | ···················· | ···················· |
| J | 36 | hCoV-19/England/ALDP-9566F1/2020\|EPI_ISL_553902\|20 | ···················· | ···················· | ····················· | ···················· | ···················· | ···················· |
| J | 37 | hCoV-19/England/ALDP-952B14/2020\|EPI_ISL_553904\|20 | ···················· | ···················· | ····················· | ···················· | ···················· | ···················· |
| J | 38 | hCoV-19/England/MILK-957890/2020\|EPI_ISL_553905\|20 | ···················· | ···················· | ····················· | ···················· | ···················· | ···················· |
| J | 39 | hCoV-19/England/ALDP-956187/2020\|EPI_ISL_553906\|20 | ···················· | ···················· | ····················· | ···················· | ···················· | ···················· |
| J | 40 | hCoV-19/England/MILK-957353/2020\|EPI_ISL_553907\|20 | ···················· | ···················· | ····················· | ···················· | ···················· | ···················· |
| J | 41 | hCoV-19/England/ALDP-955DFD/2020\|EPI_ISL_553908\|20 | ···················· | ···················· | ····················· | ···················· | ···················· | ···················· |
| J | 42 | hCoV-19/England/ALDP-9566B5/2020\|EPI_ISL_553909\|20 | ···················· | ···················· | ····················· | ···················· | ···················· | ···················· |
| J | 43 | hCoV-19/England/ALDP-955CC3/2020\|EPI_ISL_553995\|20 | ···················· | ···················· | ····················· | ···················· | ···················· | ···················· |
| J | 44 | hCoV-19/England/ALDP-95620C/2020\|EPI_ISL_553996\|20 | ···················· | ···················· | ····················· | ···················· | ···················· | ···················· |
| J | 45 | hCoV-19/England/ALDP-956460/2020\|EPI_ISL_553998\|20 | ···················· | ···················· | ····················· | ···················· | ···················· | ···················· |
| J | 46 | hCoV-19/England/ALDP-95647F/2020\|EPI_ISL_553999\|20 | ···················· | ···················· | ····················· | ···················· | ···················· | ···················· |
| J | 47 | hCoV-19/England/ALDP-6D4DDB/2020\|EPI_ISL_556100\|20 | ···················· | ···················· | ····················· | ···················· | ···················· | ···················· |
| J | 48 | hCoV-19/England/ALDP-6DA986/2020\|EPI_ISL_556101\|20 | ···················· | ···················· | ····················· | ···················· | ···················· | ···················· |
| J | 49 | hCoV-19/England/MILK-63F54F/2020\|EPI_ISL_556102\|20 | ···················· | ···················· | ····················· | ···················· | ···················· | ···················· |
| J | 50 | hCoV-19/England/ALDP-6D1406/2020\|EPI_ISL_556103\|20 | ···················· | ···················· | ····················· | ···················· | ···················· | ···················· |
| J | 51 | hCoV-19/England/MILK-95DBA0/2020\|EPI_ISL_556104\|20 | ···················· | ···················· | ····················· | ···················· | ···················· | ···················· |
| J | 52 | hCoV-19/England/ALDP-6C6A4E/2020\|EPI_ISL_556105\|20 | ···················· | ···················· | ····················· | ···················· | ···················· | ···················· |
| J | 53 | hCoV-19/England/ALDP-6DAA74/2020\|EPI_ISL_556106\|20 | ···················· | ···················· | ····················· | ···················· | ···················· | ···················· |
| J | 54 | hCoV-19/England/ALDP-6CA497/2020\|EPI_ISL_556107\|20 | ···················· | ···················· | ····················· | ···················· | ···················· | ···················· |
| J | 55 | hCoV-19/England/ALDP-6D34F5/2020\|EPI_ISL_556108\|20 | ···················· | ···················· | ····················· | ···················· | ···················· | ···················· |

| | | | | | | | |
|---|---|---|---|---|---|---|---|
| J | 56 hCoV-19/England/MILK-6422BB/2020\|EPI_ISL_556109\|20 | ···················· | ···················· | ···················· | ···················· | ···················· | ···················· |
| J | 57 hCoV-19/England/ALDP-95C066/2020\|EPI_ISL_556110\|20 | ···················· | ···················· | ···················· | ···················· | ···················· | ···················· |
| J | 58 hCoV-19/England/ALDP-94EB0C/2020\|EPI_ISL_556111\|20 | ···················· | ···················· | ···················· | ···················· | ···················· | ···················· |
| J | 59 hCoV-19/England/MILK-6438C7/2020\|EPI_ISL_556112\|20 | ···················· | ···················· | ···················· | ···················· | ···················· | ···················· |
| J | 60 hCoV-19/England/ALDP-6C6AC6/2020\|EPI_ISL_556114\|20 | ···················· | ···················· | ···················· | ···················· | ···················· | ···················· |
| J | 61 hCoV-19/England/MILK-642CCC/2020\|EPI_ISL_556116\|20 | ···················· | ···················· | ···················· | ···················· | ···················· | ···················· |
| J | 62 hCoV-19/England/ALDP-94EB75/2020\|EPI_ISL_556118\|20 | ···················· | ···················· | ···················· | ···················· | ···················· | ···················· |
| J | 63 hCoV-19/England/ALDP-6C53EA/2020\|EPI_ISL_556119\|20 | ···················· | ···················· | ···················· | ···················· | ···················· | ···················· |
| J | 64 hCoV-19/England/ALDP-6C56F0/2020\|EPI_ISL_556120\|20 | ···················· | ···················· | ···················· | ···················· | ···················· | ···················· |
| J | 65 hCoV-19/England/ALDP-6C8D9E/2020\|EPI_ISL_556121\|20 | ···················· | ···················· | ···················· | ···················· | ···················· | ···················· |
| J | 66 hCoV-19/England/ALDP-6CA109/2020\|EPI_ISL_556122\|20 | ···················· | ···················· | ···················· | ···················· | ···················· | ···················· |
| J | 67 hCoV-19/England/ALDP-635242/2020\|EPI_ISL_556125\|20 | ···················· | ···················· | ···················· | ···················· | ···················· | ···················· |
| J | 68 hCoV-19/England/MILK-630E70/2020\|EPI_ISL_556126\|20 | ···················· | ···················· | ···················· | ···················· | ···················· | ···················· |
| J | 69 hCoV-19/England/ALDP-6C5872/2020\|EPI_ISL_556127\|20 | ···················· | ···················· | ···················· | ···················· | ···················· | ···················· |
| J | 70 hCoV-19/England/ALDP-6C93E6/2020\|EPI_ISL_556128\|20 | ···················· | ···················· | ···················· | ···················· | ···················· | ···················· |
| J | 71 hCoV-19/England/ALDP-6C9A0F/2020\|EPI_ISL_556129\|20 | ···················· | ·A·················· | ···················· | ···················· | ···················· | ···················· |
| J | 72 hCoV-19/Wales/ALDP-6DA8D4/2020\|EPI_ISL_556130\|2020 | ···················· | ···················· | ···················· | ···················· | ···················· | ···················· |
| J | 73 hCoV-19/England/ALDP-94ECCD/2020\|EPI_ISL_556131\|20 | ···················· | ···················· | ···················· | ···················· | ···················· | ···················· |
| J | 74 hCoV-19/England/ALDP-6CD3B5/2020\|EPI_ISL_556132\|20 | ···················· | ···················· | ···················· | ···················· | ···················· | ···················· |
| J | 75 hCoV-19/England/ALDP-94EF2E/2020\|EPI_ISL_556136\|20 | ···················· | ···················· | ···················· | ···················· | ···················· | ···················· |
| J | 76 hCoV-19/England/MILK-632412/2020\|EPI_ISL_556138\|20 | ···················· | ···················· | ···················· | ···················· | ···················· | ···················· |
| J | 77 hCoV-19/England/MILK-642AA4/2020\|EPI_ISL_556139\|20 | ···················· | ···················· | ···················· | ···················· | ···················· | ···················· |
| J | 78 hCoV-19/England/ALDP-6D0540/2020\|EPI_ISL_556142\|20 | ···················· | ···················· | ···················· | ···················· | ···················· | ···················· |
| J | 79 hCoV-19/England/ALDP-6D1A04/2020\|EPI_ISL_556143\|20 | ···················· | ···················· | ···················· | ···················· | ···················· | ···················· |
| J | 80 hCoV-19/England/ALDP-6CCF2B/2020\|EPI_ISL_556144\|20 | ···················· | ···················· | ···················· | ···················· | ···················· | ···················· |
| J | 81 hCoV-19/England/ALDP-6D1FC9/2020\|EPI_ISL_556145\|20 | ···················· | ···················· | ···················· | ···················· | ···················· | ···················· |
| J | 82 hCoV-19/England/MILK-95D9C4/2020\|EPI_ISL_556146\|20 | ···················· | ···················· | ···················· | ···················· | ···················· | ···················· |
| J | 83 hCoV-19/England/ALDP-6D6489/2020\|EPI_ISL_556147\|20 | ···················· | ···················· | ···················· | ···················· | ···················· | ···················· |
| J | 84 hCoV-19/England/MILK-63154D/2020\|EPI_ISL_556148\|20 | ···················· | ···················· | ···················· | ···················· | ···················· | ···················· |
| J | 85 hCoV-19/England/ALDP-6C993F/2020\|EPI_ISL_556149\|20 | ···················· | ·A·················· | ···················· | ···················· | ···················· | ···················· |
| J | 86 hCoV-19/England/ALDP-6D6665/2020\|EPI_ISL_556153\|20 | ···················· | ···················· | ···················· | ···················· | ···················· | ···················· |
| J | 87 hCoV-19/England/ALDP-6397D6/2020\|EPI_ISL_556155\|20 | ···················· | ···················· | ···················· | ···················· | ···················· | ···················· |
| J | 88 hCoV-19/England/ALDP-63D462/2020\|EPI_ISL_556157\|20 | ···················· | ···················· | ···················· | ···················· | ···················· | ···················· |
| J | 89 hCoV-19/England/ALDP-94EB66/2020\|EPI_ISL_556160\|20 | ···················· | ···················· | ···················· | ···················· | ···················· | ···················· |
| J | 90 hCoV-19/England/ALDP-6D4F7B/2020\|EPI_ISL_556161\|20 | ···················· | ···················· | ···················· | ···················· | ···················· | ···················· |
| J | 91 hCoV-19/England/ALDP-6D6A0F/2020\|EPI_ISL_556162\|20 | ···················· | ···················· | ···················· | ···················· | ···················· | ···················· |
| J | 92 hCoV-19/England/MILK-641735/2020\|EPI_ISL_556163\|20 | ···················· | ···················· | ···················· | ···················· | ···················· | ···················· |
| J | 93 hCoV-19/England/MILK-642B74/2020\|EPI_ISL_556166\|20 | ···················· | ···················· | ···················· | ···················· | ···················· | ···················· |
| J | 94 hCoV-19/England/ALDP-6C44E8/2020\|EPI_ISL_556167\|20 | ···················· | ···················· | ···················· | ···················· | ···················· | ···················· |
| J | 95 hCoV-19/England/ALDP-6C66C2/2020\|EPI_ISL_556168\|20 | K··················· | ···················· | ···················· | ···················· | ···················· | ···················· |
| J | 96 hCoV-19/England/ALDP-6D2D0A/2020\|EPI_ISL_556169\|20 | ···················· | ···················· | ···················· | ···················· | ···················· | ···················· |
| J | 97 hCoV-19/England/MILK-95D7D9/2020\|EPI_ISL_556170\|20 | ···················· | ···················· | ···················· | ···················· | ···················· | ···················· |
| J | 98 hCoV-19/England/ALDP-6C5748/2020\|EPI_ISL_556171\|20 | ···················· | ···················· | ···················· | ···················· | ···················· | ···················· |
| J | 99 hCoV-19/England/MILK-62EADB/2020\|EPI_ISL_556172\|20 | ···················· | ···················· | ···················· | ···················· | ···················· | ···················· |
| J | 100 hCoV-19/England/ALDP-6CA3A9/2020\|EPI_ISL_556174\|20 | ···················· | ···················· | ···················· | ···················· | ···················· | ···················· |
| J | 101 hCoV-19/England/ALDP-6CE0AE/2020\|EPI_ISL_556175\|20 | ···················· | ···················· | ···················· | ···················· | ···················· | ···················· |
| J | 102 hCoV-19/England/MILK-95DBDD/2020\|EPI_ISL_556176\|20 | ···················· | ·A·················· | ···················· | ···················· | ···················· | ···················· |
| J | 103 hCoV-19/England/ALDP-6CF8E1/2020\|EPI_ISL_556179\|20 | ···················· | ···················· | ···················· | ···················· | ···················· | ···················· |
| J | 104 hCoV-19/England/ALDP-6D51B0/2020\|EPI_ISL_556181\|20 | ···················· | ···················· | ···················· | ···················· | ···················· | ···················· |
| J | 105 hCoV-19/England/ALDP-6C7157/2020\|EPI_ISL_556185\|20 | ···················· | ···················· | ···················· | ···················· | ···················· | ···················· |
| J | 106 hCoV-19/England/MILK-95DBFB/2020\|EPI_ISL_556187\|20 | ···················· | ···················· | ···················· | ···················· | ···················· | ···················· |
| J | 107 hCoV-19/England/MILK-873B4B/2020\|EPI_ISL_557400\|20 | ···················· | ···················· | ···················· | ···················· | ···················· | ···················· |
| J | 108 hCoV-19/England/MILK-74BA22/2020\|EPI_ISL_557401\|20 | ···················· | ···················· | ···················· | ···················· | ···················· | ···················· |
| J | 109 hCoV-19/England/ALDP-744986/2020\|EPI_ISL_557404\|20 | ···················· | ···················· | ···················· | ···················· | ···················· | ···················· |
| J | 110 hCoV-19/England/ALDP-74592B/2020\|EPI_ISL_557405\|20 | ···················· | ·R·················· | ···················· | ···················· | ···················· | ···················· |
| J | 111 hCoV-19/England/MILK-6F3CD9/2020\|EPI_ISL_557407\|20 | ···················· | ·A·················· | ···················· | ···················· | ···················· | ···················· |
| J | 112 hCoV-19/England/ALDP-74982A/2020\|EPI_ISL_557408\|20 | ···················· | ···················· | ···················· | ···················· | ···················· | ···················· |
| J | 113 hCoV-19/England/MILK-6F5A0A/2020\|EPI_ISL_557410\|20 | ···················· | ···················· | ···················· | ···················· | ···················· | ···················· |
| J | 114 hCoV-19/England/ALDP-747C3F/2020\|EPI_ISL_557411\|20 | ···················· | ···················· | ···················· | ···················· | ···················· | ···················· |
| J | 115 hCoV-19/England/MILK-866DAF/2020\|EPI_ISL_557413\|20 | ···················· | ···················· | ···················· | ···················· | ···················· | ···················· |
| J | 116 hCoV-19/England/MILK-8659E8/2020\|EPI_ISL_557416\|20 | ···················· | ···················· | ···················· | ···················· | ···················· | ···················· |
| J | 117 hCoV-19/England/ALDP-745721/2020\|EPI_ISL_557417\|20 | ···················· | ···················· | ···················· | ···················· | ···················· | ···················· |
| J | 118 hCoV-19/England/MILK-74B530/2020\|EPI_ISL_557418\|20 | ···················· | ···················· | ···················· | ···················· | ···················· | ···················· |
| J | 119 hCoV-19/England/ALDP-6FD6B3/2020\|EPI_ISL_557419\|20 | ···················· | ···················· | ···················· | ···················· | ···················· | ···················· |
| J | 120 hCoV-19/England/MILK-850A83/2020\|EPI_ISL_557421\|20 | ···················· | ···················· | ···················· | ···················· | ···················· | ···················· |
| J | 121 hCoV-19/England/ALDP-6FCBD3/2020\|EPI_ISL_557423\|20 | ···················· | ···················· | ···················· | ···················· | ···················· | ···················· |
| J | 122 hCoV-19/Wales/ALDP-6FA6D4/2020\|EPI_ISL_557425\|2020 | ···················· | ···················· | ···················· | ···················· | ···················· | ···················· |
| J | 123 hCoV-19/England/MILK-752B16/2020\|EPI_ISL_557426\|20 | ···················· | ···················· | ···················· | ···················· | ···················· | ···················· |
| J | 124 hCoV-19/England/MILK-86ECE6/2020\|EPI_ISL_557427\|20 | ···················· | ·A·················· | ···················· | ···················· | ···················· | ···················· |
| J | 125 hCoV-19/England/MILK-851C31/2020\|EPI_ISL_557429\|20 | ···················· | ···················· | ···················· | ···················· | ···················· | ···················· |
| J | 126 hCoV-19/England/MILK-850D7B/2020\|EPI_ISL_557431\|20 | ···················· | ···················· | ···················· | ···················· | ···················· | ···················· |
| J | 127 hCoV-19/England/MILK-86CE79/2020\|EPI_ISL_557432\|20 | ···················· | ···················· | ···················· | ···················· | ···················· | ···················· |
| J | 128 hCoV-19/England/ALDP-745E77/2020\|EPI_ISL_557433\|20 | ···················· | ···················· | ···················· | ···················· | ···················· | ···················· |
| J | 129 hCoV-19/England/ALDP-747E93/2020\|EPI_ISL_557434\|20 | ···················· | ···················· | ···················· | ···················· | ···················· | ···················· |
| J | 130 hCoV-19/England/MILK-74A838/2020\|EPI_ISL_557435\|20 | ···················· | ···················· | ···················· | ···················· | ···················· | ···················· |
| J | 131 hCoV-19/England/ALDP-6FCD55/2020\|EPI_ISL_557436\|20 | ···················· | ···················· | ···················· | ···················· | ···················· | ···················· |
| J | 132 hCoV-19/England/MILK-6EF371/2020\|EPI_ISL_557437\|20 | ···················· | ···················· | ···················· | ···················· | ···················· | ···················· |
| J | 133 hCoV-19/England/ALDP-749A24/2020\|EPI_ISL_557439\|20 | ···················· | ···················· | ···················· | ···················· | ···················· | ···················· |
| J | 134 hCoV-19/England/ALDP-748B8C/2020\|EPI_ISL_557440\|20 | ···················· | ···················· | ···················· | ···················· | ···················· | ···················· |
| J | 135 hCoV-19/England/MILK-870CD2/2020\|EPI_ISL_557441\|20 | ···················· | ···················· | ···················· | ···················· | ···················· | ···················· |

| | | | | | | | | |
|---|---|---|---|---|---|---|---|---|
| J | 136 | hCoV-19/England/MILK-876A1E/2020\|EPI_ISL_557442\|20 | ···················· | ···················· | ····················· | ···················· | ···················· | ···················· |
| J | 137 | hCoV-19/England/MILK-859149/2020\|EPI_ISL_557443\|20 | ···················· | ···················· | ····················· | ···················· | ···················· | ···················· |
| J | 138 | hCoV-19/England/MILK-6F9D67/2020\|EPI_ISL_557445\|20 | ···················· | ···················· | ····················· | ···················· | ···················· | ···················· |
| J | 139 | hCoV-19/England/ALDP-7442C7/2020\|EPI_ISL_557446\|20 | ···················· | ···················· | ····················· | ···················· | ···················· | ···················· |
| J | 140 | hCoV-19/England/ALDP-749505/2020\|EPI_ISL_557447\|20 | ···················· | ·A·················· | ····················· | ···················· | ···················· | ···················· |
| J | 141 | hCoV-19/England/ALDP-7474FB/2020\|EPI_ISL_557449\|20 | ···················· | ···················· | ····················· | ···················· | ···················· | ···················· |
| J | 142 | hCoV-19/England/MILK-86A725/2020\|EPI_ISL_557450\|20 | ···················· | ···················· | ····················· | ···················· | ···················· | ···················· |
| J | 143 | hCoV-19/England/MILK-854F26/2020\|EPI_ISL_557451\|20 | ···················· | ···················· | ····················· | ···················· | ···················· | ···················· |
| J | 144 | hCoV-19/England/ALDP-748418/2020\|EPI_ISL_557452\|20 | ···················· | ···················· | ····················· | ···················· | ···················· | ···················· |
| J | 145 | hCoV-19/England/ALDP-6FCAC7/2020\|EPI_ISL_557453\|20 | ···················· | ···················· | ····················· | ···················· | ···················· | ···················· |
| J | 146 | hCoV-19/England/ALDP-746FBF/2020\|EPI_ISL_557454\|20 | ···················· | ···················· | ····················· | ···················· | ···················· | ···················· |
| J | 147 | hCoV-19/England/MILK-8664D7/2020\|EPI_ISL_557455\|20 | ···················· | ···················· | ····················· | ···················· | ···················· | ···················· |
| J | 148 | hCoV-19/England/MILK-6F58B5/2020\|EPI_ISL_557456\|20 | ···················· | ···················· | ····················· | ···················· | ···················· | ···················· |
| J | 149 | hCoV-19/England/MILK-85BE22/2020\|EPI_ISL_557457\|20 | ···················· | ···················· | ····················· | ···················· | ···················· | ···················· |
| J | 150 | hCoV-19/England/MILK-74D1B2/2020\|EPI_ISL_557458\|20 | ···················· | ···················· | ····················· | ···················· | ···················· | ···················· |
| J | 151 | hCoV-19/England/MILK-870777/2020\|EPI_ISL_557459\|20 | ···················· | ···················· | ····················· | ···················· | ···················· | ···················· |
| J | 152 | hCoV-19/England/MILK-86AA2C/2020\|EPI_ISL_557460\|20 | ···················· | ···················· | ····················· | ···················· | ···················· | ···················· |
| J | 153 | hCoV-19/England/MILK-6F100C/2020\|EPI_ISL_557461\|20 | ···················· | ···················· | ····················· | ···················· | ···················· | ···················· |
| J | 154 | hCoV-19/England/MILK-863DFD/2020\|EPI_ISL_557463\|20 | ···················· | ···················· | ····················· | ···················· | ···················· | ···················· |
| J | 155 | hCoV-19/England/MILK-86BDF5/2020\|EPI_ISL_557464\|20 | ···················· | ···················· | ····················· | ···················· | ···················· | ···················· |
| J | 156 | hCoV-19/England/MILK-6F57A9/2020\|EPI_ISL_557465\|20 | ···················· | ···················· | ····················· | ···················· | ···················· | ···················· |
| J | 157 | hCoV-19/Wales/ALDP-6FA9EA/2020\|EPI_ISL_557466\|2020 | ···················· | ···················· | ····················· | ···················· | ···················· | ···················· |
| J | 158 | hCoV-19/England/MILK-74B1A5/2020\|EPI_ISL_557467\|20 | ···················· | ···················· | ····················· | ···················· | ···················· | ···················· |
| J | 159 | hCoV-19/England/ALDP-6FD7CF/2020\|EPI_ISL_557468\|20 | ···················· | ·A·················· | ····················· | ···················· | ···················· | ···················· |
| J | 160 | hCoV-19/England/ALDP-74419D/2020\|EPI_ISL_557469\|20 | ···················· | ···················· | ····················· | ···················· | ···················· | ···················· |
| J | 161 | hCoV-19/England/MILK-874E6F/2020\|EPI_ISL_557470\|20 | ···················· | ···················· | ····················· | ···················· | ···················· | ···················· |
| J | 162 | hCoV-19/England/ALDP-6FC60F/2020\|EPI_ISL_557471\|20 | ···················· | ···················· | ····················· | ···················· | ···················· | ···················· |
| J | 163 | hCoV-19/England/MILK-82A7A9/2020\|EPI_ISL_557473\|20 | ···················· | ···················· | ····················· | ···················· | ···················· | ···················· |
| J | 164 | hCoV-19/England/ALDP-745378/2020\|EPI_ISL_557476\|20 | ···················· | ···················· | ····················· | ···················· | ···················· | ···················· |
| J | 165 | hCoV-19/England/MILK-85A773/2020\|EPI_ISL_557479\|20 | ···················· | ···················· | ····················· | ···················· | ···················· | ···················· |
| J | 166 | hCoV-19/England/ALDP-744054/2020\|EPI_ISL_557481\|20 | ···················· | ···················· | ····················· | ···················· | ···················· | ···················· |
| J | 167 | hCoV-19/England/ALDP-6FD62C/2020\|EPI_ISL_557482\|20 | ···················· | ···················· | ····················· | ···················· | ···················· | ···················· |
| J | 168 | hCoV-19/England/MILK-863DDF/2020\|EPI_ISL_557483\|20 | ···················· | ···················· | ····················· | ···················· | ···················· | ···················· |
| J | 169 | hCoV-19/England/ALDP-6FD70B/2020\|EPI_ISL_557484\|20 | ···················· | ···················· | ····················· | ···················· | ···················· | ···················· |
| J | 170 | hCoV-19/England/ALDP-744449/2020\|EPI_ISL_557485\|20 | ···················· | ···················· | ····················· | ···················· | ···················· | ···················· |
| J | 171 | hCoV-19/England/MILK-867A10/2020\|EPI_ISL_557486\|20 | ···················· | ···················· | ····················· | ···················· | ···················· | ···················· |
| J | 172 | hCoV-19/England/ALDP-7478B3/2020\|EPI_ISL_557487\|20 | ···················· | ···················· | ····················· | ···················· | ···················· | ···················· |
| J | 173 | hCoV-19/England/ALDP-74431F/2020\|EPI_ISL_557488\|20 | ···················· | ···················· | ····················· | ···················· | ···················· | ···················· |
| J | 174 | hCoV-19/England/MILK-85324C/2020\|EPI_ISL_557489\|20 | ···················· | ···················· | ····················· | ···················· | ···················· | ···················· |
| J | 175 | hCoV-19/England/MILK-8603EF/2020\|EPI_ISL_557490\|20 | ···················· | ···················· | ····················· | ···················· | ···················· | ···················· |
| J | 176 | hCoV-19/England/ALDP-6FD8AE/2020\|EPI_ISL_557491\|20 | ···················· | ···················· | ····················· | ···················· | ···················· | ···················· |
| J | 177 | hCoV-19/England/MILK-751BF9/2020\|EPI_ISL_557492\|20 | ···················· | ···················· | ····················· | ···················· | ···················· | ···················· |
| J | 178 | hCoV-19/England/MILK-87213B/2020\|EPI_ISL_557495\|20 | ···················· | ······R············· | ····················· | ···················· | ···················· | ···················· |
| J | 179 | hCoV-19/England/MILK-6F65EA/2020\|EPI_ISL_557496\|20 | ···················· | ···················· | ····················· | ···················· | ···················· | ···················· |
| J | 180 | hCoV-19/England/ALDP-6FC845/2020\|EPI_ISL_557497\|20 | ···················· | ···················· | ····················· | ···················· | ···················· | ···················· |
| J | 181 | hCoV-19/England/MILK-863A23/2020\|EPI_ISL_557498\|20 | ···················· | ···················· | ····················· | ···················· | ···················· | ···················· |
| J | 182 | hCoV-19/England/ALDP-746984/2020\|EPI_ISL_557499\|20 | ···················· | ·A·················· | ····················· | ···················· | ···················· | ···················· |
| J | 183 | hCoV-19/England/ALDP-5AF226/2020\|EPI_ISL_558700\|20 | ···················· | ···················· | ····················· | ···················· | ···················· | ···················· |
| J | 184 | hCoV-19/England/ALDP-5ACD82/2020\|EPI_ISL_558703\|20 | ···················· | ···················· | ····················· | ···················· | ···················· | ···················· |
| J | 185 | hCoV-19/England/ALDP-5B7434/2020\|EPI_ISL_558705\|20 | ···················· | ···················· | ····················· | ···················· | ···················· | ···················· |
| J | 186 | hCoV-19/England/ALDP-5B713D/2020\|EPI_ISL_558706\|20 | ···················· | ···················· | ····················· | ···················· | ···················· | ···················· |
| J | 187 | hCoV-19/England/ALDP-59DE45/2020\|EPI_ISL_558707\|20 | ···················· | ···················· | ····················· | ···················· | ···················· | ···················· |
| J | 188 | hCoV-19/England/ALDP-5AA829/2020\|EPI_ISL_558709\|20 | ···················· | ···················· | ····················· | ···················· | ···················· | ···················· |
| J | 189 | hCoV-19/England/ALDP-5B4042/2020\|EPI_ISL_558711\|20 | ···················· | ···················· | ····················· | ···················· | ···················· | ···················· |
| J | 190 | hCoV-19/England/ALDP-5A2BEC/2020\|EPI_ISL_558713\|20 | ···················· | ···················· | ····················· | ···················· | ···················· | ···················· |
| J | 191 | hCoV-19/England/ALDP-5A2162/2020\|EPI_ISL_558714\|20 | ···················· | ···················· | ····················· | ···················· | ···················· | ···················· |
| J | 192 | hCoV-19/England/ALDP-5B68B1/2020\|EPI_ISL_558717\|20 | ···················· | ···················· | ····················· | ···················· | ···················· | ···················· |
| J | 193 | hCoV-19/England/ALDP-5B4938/2020\|EPI_ISL_558718\|20 | ···················· | ···················· | ····················· | ···················· | ···················· | ···················· |
| J | 194 | hCoV-19/England/ALDP-5AA610/2020\|EPI_ISL_558719\|20 | ···················· | ···················· | ····················· | ···················· | ···················· | ···················· |
| J | 195 | hCoV-19/England/ALDP-5ADA02/2020\|EPI_ISL_558721\|20 | ···················· | ···················· | ····················· | ···················· | ···················· | ···················· |
| J | 196 | hCoV-19/England/ALDP-5A0C27/2020\|EPI_ISL_558722\|20 | ···················· | ···················· | ····················· | ···················· | ···················· | ···················· |
| J | 197 | hCoV-19/England/ALDP-5B05B0/2020\|EPI_ISL_558723\|20 | ···················· | ···················· | ····················· | ···················· | ···················· | ···················· |
| J | 198 | hCoV-19/England/ALDP-5BF85E/2020\|EPI_ISL_558725\|20 | ···················· | ···················· | ····················· | ···················· | ···················· | ···················· |
| J | 199 | hCoV-19/England/ALDP-5A9E37/2020\|EPI_ISL_558726\|20 | ···················· | ···················· | ····················· | ···················· | ···················· | ···················· |
| J | 200 | hCoV-19/England/ALDP-5BA228/2020\|EPI_ISL_558727\|20 | ···················· | ···················· | ····················· | ···················· | ···················· | ···················· |
| J | 201 | hCoV-19/England/ALDP-5B8DDE/2020\|EPI_ISL_558728\|20 | ···················· | ···················· | ····················· | ···················· | ···················· | ···················· |
| J | 202 | hCoV-19/England/ALDP-5A28B8/2020\|EPI_ISL_558729\|20 | ···················· | ···················· | ····················· | ···················· | ···················· | ···················· |
| J | 203 | hCoV-19/England/MILK-5747C1/2020\|EPI_ISL_558731\|20 | ···················· | ···················· | ····················· | ···················· | ···················· | ···················· |
| J | 204 | hCoV-19/England/ALDP-5B5023/2020\|EPI_ISL_558732\|20 | ···················· | ···················· | ····················· | ···················· | ···················· | ···················· |
| J | 205 | hCoV-19/England/ALDP-5C0E94/2020\|EPI_ISL_558733\|20 | ···················· | ···················· | ····················· | ···················· | ···················· | ···················· |
| J | 206 | hCoV-19/England/ALDP-5A626B/2020\|EPI_ISL_558734\|20 | ···················· | ···················· | ····················· | ···················· | ···················· | ···················· |
| J | 207 | hCoV-19/England/ALDP-5C1C6C/2020\|EPI_ISL_558735\|20 | ···················· | ···················· | ····················· | ···················· | ···················· | ···················· |
| J | 208 | hCoV-19/England/ALDP-5AA65C/2020\|EPI_ISL_558736\|20 | ···················· | ···················· | ····················· | ···················· | ···················· | ···················· |
| J | 209 | hCoV-19/England/ALDP-5C1006/2020\|EPI_ISL_558738\|20 | ···················· | ···················· | ····················· | ···················· | ···················· | ···················· |
| J | 210 | hCoV-19/England/ALDP-5A2D40/2020\|EPI_ISL_558739\|20 | ···················· | ···················· | ····················· | ···················· | ···················· | ···················· |
| J | 211 | hCoV-19/England/ALDP-59E12D/2020\|EPI_ISL_558740\|20 | ···················· | ···················· | ····················· | ···················· | ···················· | ···················· |
| J | 212 | hCoV-19/Wales/ALDP-5A8AE9/2020\|EPI_ISL_558741\|2020 | ···················· | ···················· | ····················· | ···················· | ···················· | ···················· |
| J | 213 | hCoV-19/England/ALDP-5B5876/2020\|EPI_ISL_558742\|20 | ···················· | ·A·················· | ····················· | ···················· | ···················· | ···················· |
| J | 214 | hCoV-19/England/ALDP-5BFE5C/2020\|EPI_ISL_558743\|20 | ···················· | ···················· | ····················· | ···················· | ···················· | ···················· |
| J | 215 | hCoV-19/England/ALDP-5AB336/2020\|EPI_ISL_558744\|20 | ···················· | ···················· | ····················· | ···················· | ···················· | ···················· |

J 216 hCoV-19/England/ALDP-5C2542/2020|EPI_ISL_558745|20 ···················· ···················· ···················· ···················· ···················· ····················
J 217 hCoV-19/England/ALDP-5BCD52/2020|EPI_ISL_558746|20 ···················· ···················· ···················· ···················· ···················· ····················
J 218 hCoV-19/England/ALDP-5A3C06/2020|EPI_ISL_558747|20 ···················· ···················· ···················· ···················· ···················· ····················
J 219 hCoV-19/England/ALDP-5A30DD/2020|EPI_ISL_558748|20 ···················· ···················· ···················· ···················· ···················· ····················
J 220 hCoV-19/England/ALDP-5ACB6A/2020|EPI_ISL_558749|20 ···················· ·A·················· ···················· ···················· ···················· ····················
J 221 hCoV-19/England/ALDP-5AC292/2020|EPI_ISL_558750|20 ···················· ···················· ···················· ···················· ···················· ····················
J 222 hCoV-19/England/ALDP-5A3019/2020|EPI_ISL_558752|20 ···················· ···················· ···················· ···················· ···················· ····················
J 223 hCoV-19/Wales/ALDP-5AED35/2020|EPI_ISL_558753|2020 ···················· ···················· ···················· ···················· ···················· ····················
J 224 hCoV-19/England/ALDP-59FDCE/2020|EPI_ISL_558754|20 ···················· ···················· ···················· ···················· ···················· ····················
J 225 hCoV-19/England/ALDP-59E6E2/2020|EPI_ISL_558757|20 ···················· ···················· ···················· ···················· ···················· ····················
J 226 hCoV-19/England/ALDP-59E8DD/2020|EPI_ISL_558758|20 ···················· ···················· ···················· ···················· ···················· ····················
J 227 hCoV-19/England/ALDP-5AE96E/2020|EPI_ISL_558759|20 ···················· ···················· ···················· ···················· ···················· ····················
J 228 hCoV-19/England/ALDP-59E6F1/2020|EPI_ISL_558760|20 ···················· ···················· ···················· ···················· ···················· ····················
J 229 hCoV-19/England/ALDP-5A3B90/2020|EPI_ISL_558761|20 ···················· ···················· ···················· ···················· ···················· ····················
J 230 hCoV-19/England/ALDP-5AC186/2020|EPI_ISL_558762|20 ···················· ···················· ···················· ···················· ···················· ····················
J 231 hCoV-19/England/ALDP-5C0993/2020|EPI_ISL_558764|20 ···················· ···················· ···················· ···················· ···················· ····················
J 232 hCoV-19/England/ALDP-5AF5E1/2020|EPI_ISL_558765|20 ···················· ···················· ···················· ···················· ···················· ····················
J 233 hCoV-19/England/ALDP-5AF01D/2020|EPI_ISL_558766|20 ···················· ···················· ···················· ···················· ···················· ····················
J 234 hCoV-19/England/ALDP-5AC942/2020|EPI_ISL_558768|20 ···················· ···················· ···················· ···················· ···················· ····················
J 235 hCoV-19/England/ALDP-5A3DF4/2020|EPI_ISL_558769|20 ···················· ·A·················· ···················· ···················· ···················· ····················
J 236 hCoV-19/England/ALDP-5AB7A3/2020|EPI_ISL_558770|20 ···················· ···················· ···················· ···················· ···················· ····················
J 237 hCoV-19/England/ALDP-5B57B5/2020|EPI_ISL_558771|20 ···················· ·A·················· ···················· ···················· ···················· ····················
J 238 hCoV-19/England/ALDP-5C3909/2020|EPI_ISL_558772|20 ···················· ···················· ···················· ···················· ···················· ····················
J 239 hCoV-19/England/ALDP-5A2C07/2020|EPI_ISL_558773|20 ···················· ···················· ···················· ···················· ···················· ····················
J 240 hCoV-19/England/ALDP-5C3A7F/2020|EPI_ISL_558774|20 ···················· ···················· ···················· ···················· ···················· ····················
J 241 hCoV-19/England/ALDP-5A0629/2020|EPI_ISL_558776|20 ···················· ···················· ···················· ···················· ···················· ····················
J 242 hCoV-19/England/ALDP-5A47F5/2020|EPI_ISL_558777|20 ···················· ···················· ···················· ···················· ···················· ····················
J 243 hCoV-19/England/ALDP-5A3617/2020|EPI_ISL_558778|20 ···················· ···················· ···················· ···················· ···················· ····················
J 244 hCoV-19/England/ALDP-5A5080/2020|EPI_ISL_558779|20 ···················· ···················· ···················· ···················· ···················· ····················
J 245 hCoV-19/England/ALDP-59E5D6/2020|EPI_ISL_558780|20 ···················· ···················· ···················· ···················· ···················· ····················
J 246 hCoV-19/England/ALDP-5BC5F0/2020|EPI_ISL_558781|20 ···················· ···················· ···················· ···················· ···················· ····················
J 247 hCoV-19/England/ALDP-5ABDED/2020|EPI_ISL_558782|20 ···················· ···················· ···················· ···················· ···················· ····················
J 248 hCoV-19/Wales/ALDP-5A8E92/2020|EPI_ISL_558786|2020 ···················· ···················· ···················· ···················· ···················· ····················
J 249 hCoV-19/England/ALDP-5C3BD6/2020|EPI_ISL_558787|20 ···················· ···················· ···················· ···················· ···················· ····················
J 250 hCoV-19/England/ALDP-5BDF4C/2020|EPI_ISL_558789|20 ···················· ···················· ···················· ···················· ···················· ····················
J 251 hCoV-19/England/ALDP-5B7935/2020|EPI_ISL_558791|20 ···················· ···················· ···················· ···················· ···················· ····················
J 252 hCoV-19/England/ALDP-5AD255/2020|EPI_ISL_558793|20 ···················· ···················· ···················· ···················· ···················· ····················
J 253 hCoV-19/England/ALDP-5B94F6/2020|EPI_ISL_558795|20 ···················· ···················· ···················· ···················· ···················· ····················
J 254 hCoV-19/England/ALDP-5A4C23/2020|EPI_ISL_558797|20 ···················· ···················· ···················· ···················· ···················· ····················
J 255 hCoV-19/England/ALDP-5A32C8/2020|EPI_ISL_558798|20 ···················· ···················· ···················· ···················· ···················· ····················
J 256 hCoV-19/England/ALDP-5AF9B8/2020|EPI_ISL_558799|20 ···················· ·A·················· ···················· ···················· ···················· ····················
J 257 hCoV-19/Norway/3198/2020|EPI_ISL_549030|2020-08-20 ···················· ·A·················· ···················· ···················· ···················· ····················
J 258 hCoV-19/Norway/3199/2020|EPI_ISL_549031|2020-08-18 ···················· ···················· ···················· ···················· ···················· ····················
J 259 hCoV-19/Norway/3201/2020|EPI_ISL_549032|2020-08-08 ···················· ···················· ···················· ···················· ···················· ····················
J 260 hCoV-19/Norway/3203/2020|EPI_ISL_549033|2020-08-13 ···················· ···················· ···················· ···················· ···················· ····················
J 261 hCoV-19/Norway/3205/2020|EPI_ISL_549034|2020-08-15 ···················· ···················· ···················· ···················· ···················· ····················
J 262 hCoV-19/Norway/3207/2020|EPI_ISL_549035|2020-08-15 ···················· ···················· ···················· ···················· ···················· ····················
J 263 hCoV-19/Norway/3208/2020|EPI_ISL_549036|2020-08-26 ···················· ···················· ···················· ···················· ···················· ····················
J 264 hCoV-19/Norway/3210/2020|EPI_ISL_549037|2020-08-26 ···················· ·A·················· ···················· ···················· ···················· ····················
J 265 hCoV-19/Norway/2955/2020|EPI_ISL_549038|2020-07-29 ···················· ···················· ···················· ···················· ···················· ····················
J 266 hCoV-19/Norway/3212/2020|EPI_ISL_549039|2020-08-24 ···················· ···················· ···················· ···················· ···················· ····················
J 267 hCoV-19/Norway/3213/2020|EPI_ISL_549040|2020-08-24 ···················· ···················· ···················· ···················· ···················· ····················
J 268 hCoV-19/Norway/3214/2020|EPI_ISL_549041|2020-08-24 ···················· ·A·················· ···················· ···················· ···················· ··A·················
J 269 hCoV-19/Norway/3140/2020|EPI_ISL_549042|2020-08-08 ···················· ···················· ···················· ···················· ···················· ····················
J 270 hCoV-19/Norway/3142/2020|EPI_ISL_549043|2020-08-09 ···················· ···················· ···················· ···················· ···················· ····················
J 271 hCoV-19/Norway/3143/2020|EPI_ISL_549044|2020-08-09 ···················· ···················· ···················· ···················· ···················· ····················
J 272 hCoV-19/Norway/3145/2020|EPI_ISL_549045|2020-08-09 ···················· ···················· ···················· ···················· ···················· ····················
J 273 hCoV-19/Norway/3179/2020|EPI_ISL_549046|2020-08-07 ···················· ···················· ···················· ···················· ···················· ····················
J 274 hCoV-19/Norway/3180/2020|EPI_ISL_549047|2020-08-09 ···················· ···················· ···················· ···················· ···················· ····················
J 275 hCoV-19/Norway/2956/2020|EPI_ISL_549048|2020-07-30 ···················· ···················· ···················· ···················· ···················· ····················
J 276 hCoV-19/Norway/3165/2020|EPI_ISL_549049|2020-08-17 ···················· ·····A·············· ···················· ···················· ···················· ····················
J 277 hCoV-19/Norway/3166/2020|EPI_ISL_549050|2020-08-17 ···················· ·A·················· ···················· ···················· ···················· ····················
J 278 hCoV-19/Norway/3168/2020|EPI_ISL_549051|2020-07-27 ···················· ···················· ···················· ···················· ···················· ····················
J 279 hCoV-19/Norway/3078/2020|EPI_ISL_549052|2020-08-10 ···················· ···················· ···················· ···················· ···················· ····················
J 280 hCoV-19/Norway/3011/2020|EPI_ISL_549053|2020-08-04 ···················· ···················· ···················· ···················· ···················· ····················
J 281 hCoV-19/Norway/3020/2020|EPI_ISL_549054|2020-08-05 ···················· ···················· ···················· ···················· ···················· ····················
J 282 hCoV-19/Norway/3032/2020|EPI_ISL_549055|2020-08-06 ···················· ···················· ···················· ···················· ···················· ····················
J 283 hCoV-19/Norway/3009/2020|EPI_ISL_549056|2020-08-04 ···················· ···················· ···················· ···················· ···················· ····················
J 284 hCoV-19/Norway/3027/2020|EPI_ISL_549057|2020-08-05 ···················· ···················· ···················· ···················· ···················· ····················
J 285 hCoV-19/Norway/3030/2020|EPI_ISL_549058|2020-08-05 ···················· ···················· ···················· ···················· ···················· ····················
J 286 hCoV-19/Norway/2957/2020|EPI_ISL_549059|2020-08-01 ···················· ·A·················· ···················· ···················· ···················· ··A·················
J 287 hCoV-19/Norway/3018/2020|EPI_ISL_549060|2020-08-05 ···················· ···················· ···················· ···················· ···················· ····················
J 288 hCoV-19/Norway/3028/2020|EPI_ISL_549061|2020-08-06 ···················· ···················· ···················· ···················· ···················· ····················
J 289 hCoV-19/Norway/2997/2020|EPI_ISL_549062|2020-08-04 ···················· ···················· ···················· ···················· ···················· ····················
J 290 hCoV-19/Norway/2999/2020|EPI_ISL_549063|2020-08-01 ···················· ···················· ···················· ···················· ···················· ····················
J 291 hCoV-19/Norway/3000/2020|EPI_ISL_549064|2020-08-02 ···················· ···················· ···················· ···················· ···················· ····················
J 292 hCoV-19/Norway/3001/2020|EPI_ISL_549065|2020-08-01 ···················· ···················· ···················· ···················· ···················· ····················
J 293 hCoV-19/Norway/3002/2020|EPI_ISL_549066|2020-08-02 ···················· ···················· ···················· ···················· ···················· ····················
J 294 hCoV-19/Norway/3003/2020|EPI_ISL_549067|2020-08-03 ···················· ···················· ···················· ···················· ···················· ····················
J 295 hCoV-19/Norway/3004/2020|EPI_ISL_549068|2020-08-03 ···················· ···················· ···················· ···················· ···················· ····················

| | | | | | | | | |
|---|---|---|---|---|---|---|---|---|
| J | 296 | hCoV-19/Norway/3006/2020\|EPI_ISL_549069\|2020-07-31 | .................... | .................... | .................... | .................... | .................... | .................... |
| J | 297 | hCoV-19/Norway/2958/2020\|EPI_ISL_549070\|2020-07-29 | .................... | .................... | .................... | .................... | .................... | .................... |
| J | 298 | hCoV-19/Norway/3007/2020\|EPI_ISL_549071\|2020-07-31 | .................... | .................... | .................... | .................... | .................... | .................... |
| J | 299 | hCoV-19/Norway/3013/2020\|EPI_ISL_549072\|2020-08-04 | .................... | .................... | .................... | .................... | .................... | .................... |
| J | 300 | hCoV-19/Norway/3014/2020\|EPI_ISL_549073\|2020-08-03 | .................... | .................... | .................... | .................... | .................... | .................... |
| J | 301 | hCoV-19/Norway/3015/2020\|EPI_ISL_549074\|2020-08-04 | .................... | .................... | .................... | .................... | .................... | .................... |
| J | 302 | hCoV-19/Norway/3017/2020\|EPI_ISL_549075\|2020-08-04 | .................... | .................... | .................... | .................... | .................... | .................... |
| J | 303 | hCoV-19/Norway/3024/2020\|EPI_ISL_549076\|2020-08-05 | .................... | .................... | .................... | .................... | .................... | .................... |
| J | 304 | hCoV-19/Norway/3025/2020\|EPI_ISL_549077\|2020-08-05 | .................... | .................... | .................... | .................... | .................... | .................... |
| J | 305 | hCoV-19/Norway/3029/2020\|EPI_ISL_549078\|2020-08-05 | .................... | .................... | .................... | .................... | .................... | .................... |
| J | 306 | hCoV-19/Norway/3033/2020\|EPI_ISL_549079\|2020-08-05 | .................... | .................... | .................... | .................... | .................... | .................... |
| J | 307 | hCoV-19/Norway/3034/2020\|EPI_ISL_549080\|2020-08-05 | .................... | .................... | .................... | .................... | .................... | .................... |
| J | 308 | hCoV-19/Norway/2959/2020\|EPI_ISL_549081\|2020-07-29 | .................... | .................... | .................... | .................... | .................... | .................... |
| J | 309 | hCoV-19/Norway/3053/2020\|EPI_ISL_549082\|2020-08-07 | .................... | .................... | .................... | .................... | .................... | .................... |
| J | 310 | hCoV-19/Norway/3068/2020\|EPI_ISL_549083\|2020-08-10 | .................... | .................... | .................... | .................... | .................... | .................... |
| J | 311 | hCoV-19/Norway/3069/2020\|EPI_ISL_549084\|2020-08-10 | .................... | .................... | .................... | .................... | .................... | .................... |
| J | 312 | hCoV-19/Norway/3073/2020\|EPI_ISL_549085\|2020-08-07 | .................... | .................... | .................... | .................... | .................... | .................... |
| J | 313 | hCoV-19/Norway/3075/2020\|EPI_ISL_549086\|2020-08-07 | .................... | .................... | .................... | .................... | .................... | .................... |
| J | 314 | hCoV-19/Norway/3079/2020\|EPI_ISL_549087\|2020-08-04 | .................... | .................... | .................... | .................... | .................... | .................... |
| J | 315 | hCoV-19/Norway/3062/2020\|EPI_ISL_549088\|2020-08-08 | .................... | .................... | .................... | .................... | .................... | .................... |
| J | 316 | hCoV-19/Norway/2960/2020\|EPI_ISL_549089\|2020-07-30 | .................... | .A.................. | .................... | .................... | .................... | .................... |
| J | 317 | hCoV-19/Norway/2961/2020\|EPI_ISL_549090\|2020-08-02 | .................... | .A.................. | .................... | .................... | .................... | .................... |
| J | 318 | hCoV-19/Norway/2963/2020\|EPI_ISL_549091\|2020-07-30 | .................... | .................... | .................... | .................... | .................... | .................... |
| J | 319 | hCoV-19/Norway/2965/2020\|EPI_ISL_549092\|2020-07-26 | .................... | .................... | .................... | .................... | .................... | .................... |
| J | 320 | hCoV-19/Norway/2966/2020\|EPI_ISL_549093\|2020-07-22 | .................... | .................... | .................... | .................... | .................... | .................... |
| J | 321 | hCoV-19/Norway/2967/2020\|EPI_ISL_549094\|2020-07-24 | .................... | .................... | .................... | .................... | .................... | .................... |
| J | 322 | hCoV-19/Norway/2968/2020\|EPI_ISL_549095\|2020-07-24 | .................... | .................... | .................... | .................... | .................... | .................... |
| J | 323 | hCoV-19/Norway/2969/2020\|EPI_ISL_549096\|2020-07-24 | .................... | .................... | .................... | .................... | .................... | .................... |
| J | 324 | hCoV-19/Norway/2970/2020\|EPI_ISL_549097\|2020-07-22 | .................... | .................... | .................... | .................... | .................... | .................... |
| J | 325 | hCoV-19/Norway/2972/2020\|EPI_ISL_549098\|2020-07-24 | .................... | .................... | .................... | .................... | .................... | .................... |
| J | 326 | hCoV-19/Norway/2973/2020\|EPI_ISL_549099\|2020-07-31 | .................... | .................... | .................... | .................... | .................... | .................... |
| J | 327 | hCoV-19/England/MILK-9A5435/2020\|EPI_ISL_550013\|20 | .................... | .................... | .................... | .................... | .................... | .................... |
| J | 328 | hCoV-19/England/MILK-9A85B7/2020\|EPI_ISL_550014\|20 | .................... | .................... | .................... | .................... | .................... | .................... |
| J | 329 | hCoV-19/England/MILK-9A68CF/2020\|EPI_ISL_550015\|20 | .................... | .................... | .................... | .................... | .................... | .................... |
| J | 330 | hCoV-19/England/MILK-9A8353/2020\|EPI_ISL_550016\|20 | .................... | .A.................. | .................... | .................... | .................... | .................... |
| J | 331 | hCoV-19/Wales/MILK-9A8678/2020\|EPI_ISL_550017\|2020 | .................... | .................... | .................... | .................... | .................... | .................... |
| J | 332 | hCoV-19/England/MILK-9A66A7/2020\|EPI_ISL_550018\|20 | .................... | .................... | .................... | .................... | .................... | .................... |
| J | 333 | hCoV-19/Wales/MILK-9A53CF/2020\|EPI_ISL_550019\|2020 | .................... | .................... | .................... | .................... | .................... | .................... |
| J | 334 | hCoV-19/England/MILK-9A854E/2020\|EPI_ISL_550021\|20 | .................... | .................... | .................... | .................... | .................... | .................... |
| J | 335 | hCoV-19/England/MILK-9A5383/2020\|EPI_ISL_550022\|20 | .................... | .................... | .................... | .................... | .................... | .................... |
| J | 336 | hCoV-19/Wales/MILK-9A56D5/2020\|EPI_ISL_550023\|2020 | .................... | .................... | .................... | .................... | .................... | .................... |
| J | 337 | hCoV-19/England/MILK-9A8BA6/2020\|EPI_ISL_550024\|20 | .................... | .................... | .................... | .................... | .................... | .................... |
| J | 338 | hCoV-19/England/MILK-9A5778/2020\|EPI_ISL_550025\|20 | .................... | .................... | .................... | .................... | .................... | .................... |
| J | 339 | hCoV-19/England/MILK-9A530B/2020\|EPI_ISL_550026\|20 | .................... | .................... | .................... | .................... | .................... | .................... |
| J | 340 | hCoV-19/England/MILK-9A8DDD/2020\|EPI_ISL_550027\|20 | .................... | .................... | .................... | .................... | .................... | .................... |
| J | 341 | hCoV-19/Wales/MILK-9A51C5/2020\|EPI_ISL_550030\|2020 | .................... | .................... | .................... | .................... | .................... | .................... |
| J | 342 | hCoV-19/England/MILK-9A55D8/2020\|EPI_ISL_550031\|20 | .................... | .................... | .................... | .................... | .....T.............. | .................... |
| J | 343 | hCoV-19/England/MILK-9A507D/2020\|EPI_ISL_550032\|20 | .................... | .................... | .................... | .................... | .................... | .................... |
| J | 344 | hCoV-19/England/MILK-9A861E/2020\|EPI_ISL_550033\|20 | .................... | .................... | .................... | .................... | .................... | .................... |
| J | 345 | hCoV-19/England/MILK-9A55E7/2020\|EPI_ISL_550034\|20 | .................... | .A.................. | .................... | .................... | .................... | ..A................. |
| J | 346 | hCoV-19/England/MILK-9A55BA/2020\|EPI_ISL_550035\|20 | .................... | .................... | .................... | .................... | .....T.............. | .................... |
| J | 347 | hCoV-19/Wales/MILK-9A5532/2020\|EPI_ISL_550036\|2020 | .................... | .................... | .................... | .................... | .................... | .................... |
| J | 348 | hCoV-19/Wales/MILK-9A8CB2/2020\|EPI_ISL_550037\|2020 | .................... | .................... | .................... | .................... | .................... | .................... |
| J | 349 | hCoV-19/England/MILK-9A5620/2020\|EPI_ISL_550038\|20 | .................... | .................... | .................... | .................... | .................... | .................... |
| J | 350 | hCoV-19/England/MILK-9A6689/2020\|EPI_ISL_550039\|20 | .................... | .................... | .................... | .................... | .................... | .................... |
| J | 351 | hCoV-19/England/MILK-9A6AD8/2020\|EPI_ISL_550040\|20 | .................... | .................... | .................... | .................... | .................... | .................... |
| J | 352 | hCoV-19/Wales/MILK-9A574B/2020\|EPI_ISL_550041\|2020 | .................... | .................... | .................... | .................... | .................... | .................... |
| J | 353 | hCoV-19/England/MILK-9A5699/2020\|EPI_ISL_550042\|20 | .................... | .................... | .................... | .................... | .................... | .................... |
| J | 354 | hCoV-19/England/MILK-9A563F/2020\|EPI_ISL_550043\|20 | .................... | .................... | .................... | .................... | .................... | .................... |
| J | 355 | hCoV-19/England/MILK-9A6C96/2020\|EPI_ISL_550044\|20 | .................... | .................... | .................... | .................... | .................... | .................... |
| J | 356 | hCoV-19/England/MILK-9A5471/2020\|EPI_ISL_550045\|20 | .................... | .................... | .................... | .................... | .................... | .................... |
| J | 357 | hCoV-19/Wales/MILK-9A8687/2020\|EPI_ISL_550046\|2020 | .................... | .................... | .................... | .................... | .................... | .................... |
| J | 358 | hCoV-19/England/MILK-9A6C78/2020\|EPI_ISL_550047\|20 | .................... | .................... | .................... | .................... | .................... | .................... |
| J | 359 | hCoV-19/England/MILK-9A6D1B/2020\|EPI_ISL_550048\|20 | .................... | .................... | .................... | .................... | .................... | .................... |
| J | 360 | hCoV-19/England/MILK-9A8B97/2020\|EPI_ISL_550049\|20 | .................... | .................... | .................... | .................... | .................... | .................... |
| J | 361 | hCoV-19/England/MILK-9A8C85/2020\|EPI_ISL_550050\|20 | .................... | .................... | .................... | .................... | .................... | .................... |
| J | 362 | hCoV-19/England/MILK-9A50B9/2020\|EPI_ISL_550051\|20 | .................... | .A.................. | .................... | .................... | .................... | ..A................. |
| J | 363 | hCoV-19/England/MILK-9A8D19/2020\|EPI_ISL_550052\|20 | .................... | .................... | .................... | .................... | .................... | .................... |
| J | 364 | hCoV-19/England/MILK-9A8CA3/2020\|EPI_ISL_550053\|20 | .................... | ......A............. | .................... | .................... | .................... | .................... |
| J | 365 | hCoV-19/England/MILK-9A5286/2020\|EPI_ISL_550054\|20 | .................... | .................... | .................... | .................... | .................... | .................... |
| J | 366 | hCoV-19/England/MILK-9A6D39/2020\|EPI_ISL_550055\|20 | .................... | .................... | .................... | .................... | .................... | .................... |
| J | 367 | hCoV-19/England/MILK-9A6DC0/2020\|EPI_ISL_550056\|20 | .................... | .................... | .................... | .................... | .................... | .................... |
| J | 368 | hCoV-19/England/MILK-9A8DBF/2020\|EPI_ISL_550057\|20 | .................... | .................... | .................... | .................... | .................... | .................... |
| J | 369 | hCoV-19/England/MILK-9A67E0/2020\|EPI_ISL_550058\|20 | .................... | .................... | .................... | .................... | .................... | .................... |
| J | 370 | hCoV-19/England/MILK-9A5295/2020\|EPI_ISL_550059\|20 | .................... | .................... | .................... | .................... | .................... | .................... |
| J | 371 | hCoV-19/England/MILK-9A6768/2020\|EPI_ISL_550060\|20 | .................... | .................... | .................... | .................... | .................... | .................... |
| J | 372 | hCoV-19/England/MILK-9A8D37/2020\|EPI_ISL_550061\|20 | .................... | .................... | .................... | .................... | .................... | .................... |
| J | 373 | hCoV-19/England/MILK-9A67B3/2020\|EPI_ISL_550062\|20 | .................... | .................... | .................... | .................... | .................... | .................... |
| J | 374 | hCoV-19/England/MILK-9A8BB5/2020\|EPI_ISL_550063\|20 | .................... | .................... | .................... | .................... | .................... | .................... |
| J | 375 | hCoV-19/England/MILK-9A508C/2020\|EPI_ISL_550064\|20 | .................... | .................... | .................... | .................... | .................... | .................... |

| | | | | | | | |
|---|---|---|---|---|---|---|---|
| J | 376 hCoV-19/England/MILK-9A6ABA/2020\|EPI_ISL_550065\|20 | ···················· | ···················· | ···················· | ···················· | ···················· | ···················· |
| J | 377 hCoV-19/England/MILK-9A6838/2020\|EPI_ISL_550066\|20 | ···················· | ···················· | ···················· | ···················· | ···················· | ···················· |
| J | 378 hCoV-19/England/MILK-9A8D46/2020\|EPI_ISL_550067\|20 | ···················· | ···················· | ···················· | ···················· | ···················· | ···················· |
| J | 379 hCoV-19/England/MILK-9A85E4/2020\|EPI_ISL_550068\|20 | ···················· | ···················· | ···················· | ···················· | ···················· | ···················· |
| J | 380 hCoV-19/England/MILK-9A6B7B/2020\|EPI_ISL_550070\|20 | ···················· | ·A·······A·········· | ···················· | ···················· | ···················· | ···················· |
| J | 381 hCoV-19/Wales/MILK-9A50AA/2020\|EPI_ISL_550071\|2020 | ···················· | ···················· | ···················· | ···················· | ···················· | ···················· |
| J | 382 hCoV-19/England/MILK-9A8AD6/2020\|EPI_ISL_550073\|20 | ···················· | ···················· | ···················· | ···················· | ···················· | ···················· |
| J | 383 hCoV-19/England/MILK-9A5514/2020\|EPI_ISL_550074\|20 | ···················· | ···················· | ···················· | ···················· | ···················· | ···················· |
| J | 384 hCoV-19/England/MILK-9A6829/2020\|EPI_ISL_550075\|20 | ···················· | ···················· | ···················· | ···················· | ···················· | ···················· |
| J | 385 hCoV-19/England/MILK-9A8C94/2020\|EPI_ISL_550077\|20 | ···················· | ···················· | ···················· | ···················· | ···················· | ···················· |
| J | 386 hCoV-19/England/MILK-9A56C6/2020\|EPI_ISL_550078\|20 | ···················· | ····A··············· | ···················· | ···················· | ···················· | ···················· |
| J | 387 hCoV-19/England/MILK-9A8DCE/2020\|EPI_ISL_550079\|20 | ···················· | ···················· | ···················· | ···················· | ···················· | ···················· |
| J | 388 hCoV-19/England/MILK-9A856C/2020\|EPI_ISL_550080\|20 | ···················· | ···················· | ···················· | ···················· | ···················· | ···················· |
| J | 389 hCoV-19/England/MILK-9A524A/2020\|EPI_ISL_550081\|20 | ···················· | ···················· | ···················· | ···················· | ······T············· | ···················· |
| J | 390 hCoV-19/England/MILK-9A847E/2020\|EPI_ISL_550082\|20 | ···················· | ···················· | ···················· | ···················· | ···················· | ···················· |
| J | 391 hCoV-19/England/MILK-9A6A6F/2020\|EPI_ISL_550083\|20 | ···················· | ···················· | ···················· | ···················· | ···················· | ···················· |
| J | 392 hCoV-19/England/MILK-9A6ECD/2020\|EPI_ISL_550084\|20 | ···················· | ···················· | ···················· | ···················· | ···················· | ···················· |
| J | 393 hCoV-19/England/MILK-9A50F5/2020\|EPI_ISL_550085\|20 | ···················· | ···················· | ···················· | ···················· | ···················· | ···················· |
| J | 394 hCoV-19/England/MILK-9A6B5D/2020\|EPI_ISL_550086\|20 | ···················· | ···················· | ···················· | ···················· | ···················· | ···················· |
| J | 395 hCoV-19/England/MILK-9A513E/2020\|EPI_ISL_550088\|20 | ···················· | ·A·················· | ···················· | ···················· | ···················· | ···A················ |
| J | 396 hCoV-19/England/MILK-9A8DEC/2020\|EPI_ISL_550089\|20 | ···················· | ···················· | ···················· | ···················· | ···················· | ···················· |
| J | 397 hCoV-19/England/ALDP-951B51/2020\|EPI_ISL_551344\|20 | ···················· | ·A·················· | ···················· | ···················· | ···················· | ···················· |
| J | 398 hCoV-19/England/ALDP-951E1C/2020\|EPI_ISL_551345\|20 | ···················· | ···················· | ···················· | ···················· | ···················· | ···················· |
| J | 399 hCoV-19/England/ALDP-951C30/2020\|EPI_ISL_551346\|20 | ···················· | ···················· | ···················· | ···················· | ···················· | ···················· |
| J | 400 hCoV-19/England/ALDP-951FBF/2020\|EPI_ISL_551347\|20 | ···················· | ···················· | ···················· | ···················· | ···················· | ···················· |
| J | 401 hCoV-19/England/ALDP-95232B/2020\|EPI_ISL_551348\|20 | ···················· | ···················· | ···················· | ···················· | ···················· | ···················· |
| J | 402 hCoV-19/England/ALDP-9522D3/2020\|EPI_ISL_551350\|20 | ···················· | ···················· | ···················· | ···················· | ···················· | ···················· |
| J | 403 hCoV-19/England/ALDP-951F37/2020\|EPI_ISL_551351\|20 | ···················· | ···················· | ···················· | ···················· | ···················· | ···················· |
| J | 404 hCoV-19/England/ALDP-936615/2020\|EPI_ISL_551354\|20 | ···················· | ···················· | ···················· | ···················· | ···················· | ···················· |
| J | 405 hCoV-19/England/ALDP-9389FC/2020\|EPI_ISL_551355\|20 | ···················· | ···················· | ···················· | ···················· | ···················· | ···················· |
| J | 406 hCoV-19/England/ALDP-905970/2020\|EPI_ISL_551356\|20 | ···················· | ···················· | ···················· | ···················· | ···················· | ···················· |
| J | 407 hCoV-19/England/ALDP-90B8D7/2020\|EPI_ISL_551357\|20 | ···················· | ···················· | ···················· | ···················· | ···················· | ···················· |
| J | 408 hCoV-19/England/ALDP-90655D/2020\|EPI_ISL_551359\|20 | ···················· | ···················· | ···················· | ···················· | ···················· | ···················· |
| J | 409 hCoV-19/England/ALDP-9387E3/2020\|EPI_ISL_551360\|20 | ···················· | ···················· | ···················· | ···················· | ···················· | ···················· |
| J | 410 hCoV-19/England/ALDP-90EC14/2020\|EPI_ISL_551361\|20 | ···················· | ···················· | ···················· | ···················· | ···················· | ···················· |
| J | 411 hCoV-19/England/ALDP-932734/2020\|EPI_ISL_551362\|20 | ···················· | ·A·················· | ···················· | ···················· | ···················· | ···················· |
| J | 412 hCoV-19/England/ALDP-9065A8/2020\|EPI_ISL_551363\|20 | ···················· | ···················· | ···················· | ···················· | ···················· | ···················· |
| J | 413 hCoV-19/England/ALDP-923DBC/2020\|EPI_ISL_551364\|20 | ···················· | ···················· | ···················· | ···················· | ···················· | ···················· |
| J | 414 hCoV-19/England/ALDP-9051F0/2020\|EPI_ISL_551365\|20 | ···················· | ···················· | ···················· | ···················· | ···················· | ···················· |
| J | 415 hCoV-19/England/ALDP-900F55/2020\|EPI_ISL_551367\|20 | ···················· | ···················· | ···················· | ···················· | ···················· | ···················· |
| J | 416 hCoV-19/England/ALDP-8FEC72/2020\|EPI_ISL_551368\|20 | ···················· | ···················· | ···················· | ···················· | ···················· | ···················· |
| J | 417 hCoV-19/England/ALDP-908834/2020\|EPI_ISL_551369\|20 | ···················· | ···················· | ···················· | ···················· | ···················· | ···················· |
| J | 418 hCoV-19/England/ALDP-90C78E/2020\|EPI_ISL_551370\|20 | ···················· | ···················· | ···················· | ···················· | ···················· | ···················· |
| J | 419 hCoV-19/England/ALDP-92ACA9/2020\|EPI_ISL_551371\|20 | ···················· | ···················· | ···················· | ···················· | ···················· | ···················· |
| J | 420 hCoV-19/England/ALDP-9101A7/2020\|EPI_ISL_551372\|20 | ···················· | ·A·················· | ···················· | ···················· | ···················· | ···················· |
| J | 421 hCoV-19/England/ALDP-92BD2D/2020\|EPI_ISL_551373\|20 | ···················· | ···················· | ···················· | ···················· | ···················· | ···················· |
| J | 422 hCoV-19/England/ALDP-8FC1EE/2020\|EPI_ISL_551375\|20 | ···················· | ···················· | ···················· | ···················· | ···················· | ···················· |
| J | 423 hCoV-19/England/ALDP-90218A/2020\|EPI_ISL_551376\|20 | ···················· | ···················· | ···················· | ···················· | ···················· | ···················· |
| J | 424 hCoV-19/England/ALDP-923772/2020\|EPI_ISL_551377\|20 | ···················· | ···················· | ···················· | ···················· | ···················· | ···················· |
| J | 425 hCoV-19/England/ALDP-8FBE60/2020\|EPI_ISL_551378\|20 | ···················· | ···················· | ···················· | ···················· | ···················· | ···················· |
| J | 426 hCoV-19/England/ALDP-9387F2/2020\|EPI_ISL_551379\|20 | ···················· | ···················· | ···················· | ···················· | ···················· | ···················· |
| J | 427 hCoV-19/England/ALDP-90D6AE/2020\|EPI_ISL_551381\|20 | ···················· | ···················· | ···················· | ···················· | ···················· | ···················· |
| J | 428 hCoV-19/England/ALDP-90604D/2020\|EPI_ISL_551382\|20 | ···················· | ···················· | ···················· | ···················· | ···················· | ···················· |
| J | 429 hCoV-19/England/ALDP-90CF86/2020\|EPI_ISL_551383\|20 | ···················· | ···················· | ···················· | ···················· | ···················· | ···················· |
| J | 430 hCoV-19/England/ALDP-910B7C/2020\|EPI_ISL_551385\|20 | ···················· | ·A·················· | ···················· | ···················· | ···················· | ···················· |
| J | 431 hCoV-19/England/ALDP-92EF7F/2020\|EPI_ISL_551386\|20 | ···················· | ···················· | ···················· | ···················· | ···················· | ···················· |
| J | 432 hCoV-19/England/ALDP-92AEFE/2020\|EPI_ISL_551387\|20 | ···················· | ···················· | ···················· | ···················· | ···················· | ···················· |
| J | 433 hCoV-19/England/ALDP-8FB2BF/2020\|EPI_ISL_551388\|20 | ···················· | ···················· | ···················· | ···················· | ···················· | ···················· |
| J | 434 hCoV-19/England/ALDP-92B112/2020\|EPI_ISL_551389\|20 | ···················· | ···················· | ···················· | ···················· | ···················· | ···················· |
| J | 435 hCoV-19/England/ALDP-92BB50/2020\|EPI_ISL_551390\|20 | ···················· | ···················· | ···················· | ···················· | ···················· | ···················· |
| J | 436 hCoV-19/England/ALDP-90B488/2020\|EPI_ISL_551391\|20 | ···················· | ···················· | ···················· | ···················· | ···················· | ···················· |
| J | 437 hCoV-19/England/ALDP-92DD49/2020\|EPI_ISL_551392\|20 | ···················· | ···················· | ···················· | ···················· | ···················· | ···················· |
| J | 438 hCoV-19/England/ALDP-91B607/2020\|EPI_ISL_551393\|20 | ···················· | ···················· | ···················· | ···················· | ···················· | ···················· |
| J | 439 hCoV-19/England/ALDP-90C7BB/2020\|EPI_ISL_551394\|20 | ···················· | ·A·················· | ···················· | ···················· | ···················· | ···················· |
| J | 440 hCoV-19/England/ALDP-937B60/2020\|EPI_ISL_551395\|20 | ···················· | ···················· | ···················· | ···················· | ···················· | ···················· |
| J | 441 hCoV-19/England/ALDP-8FD930/2020\|EPI_ISL_551396\|20 | ···················· | ···················· | ···················· | ···················· | ···················· | ···················· |
| J | 442 hCoV-19/England/ALDP-932813/2020\|EPI_ISL_551397\|20 | ···················· | ···················· | ···················· | ···················· | ···················· | ···················· |
| J | 443 hCoV-19/England/ALDP-91012F/2020\|EPI_ISL_551399\|20 | ···················· | ···················· | ···················· | ···················· | ···················· | ···················· |
| J | 444 hCoV-19/England/MILK-975C63/2020\|EPI_ISL_552601\|20 | ···················· | ···················· | ···················· | ···················· | ···················· | ···················· |
| J | 445 hCoV-19/England/MILK-97598A/2020\|EPI_ISL_552602\|20 | ···················· | ···················· | ···················· | ···················· | ···················· | ···················· |
| J | 446 hCoV-19/England/MILK-97596C/2020\|EPI_ISL_552603\|20 | ···················· | ···················· | ···················· | ···················· | ···················· | ···················· |
| J | 447 hCoV-19/England/MILK-9756ED/2020\|EPI_ISL_552604\|20 | ···················· | ···················· | ···················· | ···················· | ···················· | ···················· |
| J | 448 hCoV-19/England/MILK-975A96/2020\|EPI_ISL_552606\|20 | ···················· | ···················· | ···················· | ···················· | ···················· | ···················· |
| J | 449 hCoV-19/England/ALDP-982B57/2020\|EPI_ISL_552608\|20 | ···················· | ···················· | ···················· | ···················· | ···················· | ···················· |
| J | 450 hCoV-19/England/MILK-974F7A/2020\|EPI_ISL_552609\|20 | ···················· | ···················· | ···················· | ···················· | ···················· | ···················· |
| J | 451 hCoV-19/England/MILK-975489/2020\|EPI_ISL_552613\|20 | ···················· | ···················· | ···················· | ···················· | ···················· | ···················· |
| J | 452 hCoV-19/England/ALDP-9814F3/2020\|EPI_ISL_552614\|20 | ···················· | ···················· | ···················· | ···················· | ···················· | ···················· |
| J | 453 hCoV-19/England/ALDP-982C18/2020\|EPI_ISL_552617\|20 | ···················· | ···················· | ···················· | ···················· | ···················· | ···················· |
| J | 454 hCoV-19/England/MILK-9755C2/2020\|EPI_ISL_552619\|20 | ···················· | ···················· | ···················· | ···················· | ···················· | ···················· |
| J | 455 hCoV-19/England/MILK-974E9B/2020\|EPI_ISL_552675\|20 | ···················· | ···················· | ···················· | ···················· | ···················· | ···················· |

| | | | | | | | | |
|---|---|---|---|---|---|---|---|---|
| J | 456 | hCoV-19/England/MILK-975225/2020\|EPI_ISL_552676\|20 | ···················· | ····A··············· | ···················· | ···················· | ···················· | ···················· |
| J | 457 | hCoV-19/England/ALDP-982D51/2020\|EPI_ISL_552679\|20 | ···················· | ···················· | ···················· | ···················· | ···················· | ···················· |
| J | 458 | hCoV-19/England/MILK-975067/2020\|EPI_ISL_552680\|20 | ···················· | ···················· | ···················· | ···················· | ···················· | ···················· |
| J | 459 | hCoV-19/England/ALDP-982823/2020\|EPI_ISL_552681\|20 | ···················· | ···················· | ···················· | ···················· | ···················· | ···················· |
| J | 460 | hCoV-19/England/MILK-975058/2020\|EPI_ISL_552682\|20 | ···················· | ···················· | ···················· | ···················· | ···················· | ···················· |
| J | 461 | hCoV-19/England/MILK-974F2F/2020\|EPI_ISL_552684\|20 | ···················· | C··················· | ···················· | ···················· | ···················· | ···················· |
| J | 462 | hCoV-19/England/MILK-9758F6/2020\|EPI_ISL_552685\|20 | ···················· | ···················· | ···················· | ···················· | ···················· | ···················· |
| J | 463 | hCoV-19/England/ALDP-982B2A/2020\|EPI_ISL_552686\|20 | ···················· | ···················· | ···················· | ···················· | ···················· | ···················· |
| J | 464 | hCoV-19/England/MILK-974C55/2020\|EPI_ISL_552688\|20 | ···················· | ···················· | ···················· | ···················· | ···················· | ···················· |
| J | 465 | hCoV-19/England/ALDP-98296C/2020\|EPI_ISL_552689\|20 | ···················· | ···················· | ···················· | ···················· | ···················· | ···················· |
| J | 466 | hCoV-19/England/MILK-9756FC/2020\|EPI_ISL_552690\|20 | ···················· | ·A·················· | ···················· | ···················· | ···················· | ··A················· |
| J | 467 | hCoV-19/England/MILK-975814/2020\|EPI_ISL_552691\|20 | ···················· | ···················· | ···················· | ···················· | ···················· | ···················· |
| J | 468 | hCoV-19/England/MILK-97589C/2020\|EPI_ISL_552692\|20 | ···················· | ···················· | ···················· | ···················· | ···················· | ···················· |
| J | 469 | hCoV-19/England/MILK-974E6E/2020\|EPI_ISL_552693\|20 | ···················· | ···················· | ···················· | ···················· | ···················· | ···················· |
| J | 470 | hCoV-19/England/MILK-974CFB/2020\|EPI_ISL_552694\|20 | ···················· | ···················· | ···················· | ···················· | ···················· | ···················· |
| J | 471 | hCoV-19/England/MILK-975C36/2020\|EPI_ISL_552695\|20 | ···················· | ···················· | ···················· | ···················· | ···················· | ···················· |
| J | 472 | hCoV-19/England/MILK-975049/2020\|EPI_ISL_552696\|20 | ···················· | ···················· | ···················· | ···················· | ···················· | ···················· |
| J | 473 | hCoV-19/England/MILK-9756B0/2020\|EPI_ISL_552697\|20 | ···················· | ···················· | ···················· | ···················· | ···················· | ···················· |
| J | 474 | hCoV-19/England/MILK-9757DB/2020\|EPI_ISL_552698\|20 | ···················· | ···················· | ···················· | ···················· | ···················· | ···················· |
| J | 475 | hCoV-19/England/ALDP-9829D5/2020\|EPI_ISL_552699\|20 | ···················· | ···················· | ···················· | ···················· | ···················· | ···················· |
| J | 476 | hCoV-19/England/ALDP-95654F/2020\|EPI_ISL_553910\|20 | ···················· | ···················· | ···················· | ···················· | ···················· | ···················· |
| J | 477 | hCoV-19/England/ALDP-955B99/2020\|EPI_ISL_553911\|20 | ···················· | ···················· | ···················· | ···················· | ···················· | ···················· |
| J | 478 | hCoV-19/England/ALDP-955D93/2020\|EPI_ISL_553912\|20 | ···················· | ···················· | ···················· | ···················· | ···················· | ···················· |
| J | 479 | hCoV-19/England/ALDP-955DC0/2020\|EPI_ISL_553915\|20 | ···················· | ···················· | ···················· | ···················· | ···················· | ···················· |
| J | 480 | hCoV-19/England/ALDP-952947/2020\|EPI_ISL_553916\|20 | ···················· | ···················· | ···················· | ···················· | ···················· | ···················· |
| J | 481 | hCoV-19/England/ALDP-9529B0/2020\|EPI_ISL_553917\|20 | ···················· | ···················· | ···················· | ···················· | ···················· | ···················· |
| J | 482 | hCoV-19/England/ALDP-9561D2/2020\|EPI_ISL_553918\|20 | ···················· | ···················· | ···················· | ···················· | ···················· | ···················· |
| J | 483 | hCoV-19/England/ALDP-955F9D/2020\|EPI_ISL_553919\|20 | ···················· | ···················· | ···················· | ···················· | ···················· | ···················· |
| J | 484 | hCoV-19/England/ALDP-952701/2020\|EPI_ISL_553921\|20 | ···················· | ···················· | ···················· | ···················· | ···················· | ···················· |
| J | 485 | hCoV-19/England/ALDP-952ABD/2020\|EPI_ISL_553922\|20 | ···················· | ···················· | ···················· | ···················· | ···················· | ···················· |
| J | 486 | hCoV-19/England/ALDP-952ACC/2020\|EPI_ISL_553923\|20 | ···················· | ···················· | ···················· | ···················· | ···················· | ···················· |
| J | 487 | hCoV-19/England/ALDP-956318/2020\|EPI_ISL_553925\|20 | ···················· | ···················· | ···················· | ···················· | ···················· | ···················· |
| J | 488 | hCoV-19/England/ALDP-955F15/2020\|EPI_ISL_553926\|20 | ···················· | ···················· | ···················· | ···················· | ···················· | ···················· |
| J | 489 | hCoV-19/England/ALDP-9526F5/2020\|EPI_ISL_553928\|20 | ···················· | ···················· | ···················· | ···················· | ···················· | ···················· |
| J | 490 | hCoV-19/Wales/MILK-9575D5/2020\|EPI_ISL_553929\|2020 | ···················· | ···················· | ···················· | ···················· | ···················· | ···················· |
| J | 491 | hCoV-19/England/ALDP-95266E/2020\|EPI_ISL_553930\|20 | ···················· | ···················· | ···················· | ···················· | ···················· | ···················· |
| J | 492 | hCoV-19/England/ALDP-955D66/2020\|EPI_ISL_553931\|20 | ···················· | ···················· | ···················· | ···················· | ···················· | ···················· |
| J | 493 | hCoV-19/England/ALDP-952974/2020\|EPI_ISL_553932\|20 | ···················· | ···················· | ···················· | ···················· | ···················· | ···················· |
| J | 494 | hCoV-19/England/MILK-957678/2020\|EPI_ISL_553934\|20 | ···················· | ···················· | ···················· | ···················· | ···················· | ···················· |
| J | 495 | hCoV-19/England/ALDP-9525AD/2020\|EPI_ISL_553935\|20 | ···················· | ···················· | ···················· | ···················· | ···················· | ···················· |
| J | 496 | hCoV-19/England/ALDP-9560F3/2020\|EPI_ISL_553936\|20 | ···················· | ···················· | ···················· | ···················· | ···················· | ···················· |
| J | 497 | hCoV-19/England/ALDP-9563FA/2020\|EPI_ISL_553937\|20 | ···················· | ···················· | ···················· | ···················· | ···················· | ···················· |
| J | 498 | hCoV-19/England/ALDP-9566A6/2020\|EPI_ISL_553938\|20 | ···················· | ···················· | ···················· | ···················· | ···················· | ···················· |
| J | 499 | hCoV-19/England/MILK-95754E/2020\|EPI_ISL_553939\|20 | ···················· | ···················· | ···················· | ···················· | ···················· | ···················· |
| J | 500 | hCoV-19/England/ALDP-9528E0/2020\|EPI_ISL_553947\|20 | ···················· | ···················· | ···················· | ···················· | ···················· | ···················· |
| J | 501 | hCoV-19/England/ALDP-955CE1/2020\|EPI_ISL_553948\|20 | ···················· | ···················· | ···················· | ···················· | ···················· | ···················· |
| J | 502 | hCoV-19/England/ALDP-95613C/2020\|EPI_ISL_553949\|20 | ···················· | ···················· | ···················· | ···················· | ···················· | ···················· |
| J | 503 | hCoV-19/England/ALDP-952789/2020\|EPI_ISL_553950\|20 | ···················· | ···················· | ···················· | ···················· | ···················· | ···················· |
| J | 504 | hCoV-19/England/ALDP-6353A9/2020\|EPI_ISL_556190\|20 | ···················· | ···················· | ···················· | ···················· | ···················· | ···················· |
| J | 505 | hCoV-19/England/ALDP-6C90C1/2020\|EPI_ISL_556193\|20 | ···················· | ···················· | ···················· | ···················· | ···················· | ···················· |
| J | 506 | hCoV-19/England/ALDP-6D03EC/2020\|EPI_ISL_556194\|20 | ···················· | ···················· | ···················· | ···················· | ···················· | ···················· |
| J | 507 | hCoV-19/England/MILK-631F9A/2020\|EPI_ISL_556195\|20 | ···················· | ···················· | ···················· | ···················· | ···················· | ···················· |
| J | 508 | hCoV-19/England/ALDP-635251/2020\|EPI_ISL_556196\|20 | ···················· | ···················· | ···················· | ···················· | ···················· | ···················· |
| J | 509 | hCoV-19/Wales/MILK-6412E9/2020\|EPI_ISL_556197\|2020 | ···················· | ···················· | ···················· | ···················· | ···················· | ···················· |
| J | 510 | hCoV-19/Wales/MILK-632922/2020\|EPI_ISL_556198\|2020 | ···················· | ···················· | ···················· | ···················· | ···················· | ···················· |
| J | 511 | hCoV-19/England/MILK-95DBEC/2020\|EPI_ISL_556199\|20 | ···················· | ···················· | ···················· | ···················· | ···················· | ···················· |
| J | 512 | hCoV-19/England/ALDP-5C18E0/2020\|EPI_ISL_558801\|20 | ···················· | ···················· | ···················· | ···················· | ···················· | ···················· |
| J | 513 | hCoV-19/Singapore/992/2020\|EPI_ISL_549000\|2020-03- | ···················· | ···················· | ···················· | ···················· | ···················· | ···················· |
| J | 514 | hCoV-19/Singapore/993/2020\|EPI_ISL_549001\|2020-03- | ···················· | ···················· | ···················· | ···················· | ···················· | ···················· |
| J | 515 | hCoV-19/Singapore/994/2020\|EPI_ISL_549002\|2020-03- | ···················· | ···················· | ···················· | ···················· | ···················· | ···················· |
| J | 516 | hCoV-19/Singapore/995/2020\|EPI_ISL_549003\|2020-03- | ···················· | ···················· | ···················· | ···················· | ···················· | ···················· |
| J | 517 | hCoV-19/Singapore/996/2020\|EPI_ISL_549004\|2020-03- | ···················· | ···················· | ···················· | ···················· | ···················· | ···················· |
| J | 518 | hCoV-19/Singapore/997/2020\|EPI_ISL_549005\|2020-03- | ···················· | ···················· | ···················· | ···················· | ···················· | ···················· |
| J | 519 | hCoV-19/Wales/MILK-9A67A4/2020\|EPI_ISL_550000\|2020 | ···················· | ···················· | ···················· | ···················· | ···················· | ···················· |
| J | 520 | hCoV-19/England/MILK-9A6D57/2020\|EPI_ISL_550001\|20 | ···················· | ···················· | ···················· | ···················· | ···················· | ···················· |
| J | 521 | hCoV-19/England/ALDP-951B9D/2020\|EPI_ISL_551301\|20 | ···················· | ···················· | ···················· | ···················· | ···················· | ···················· |
| J | 522 | hCoV-19/England/ALDP-951F46/2020\|EPI_ISL_551302\|20 | ···················· | ···················· | ···················· | ···················· | ···················· | ···················· |
| J | 523 | hCoV-19/England/ALDP-951D6A/2020\|EPI_ISL_551304\|20 | ···················· | ·A·················· | ···················· | ···················· | ···················· | ···················· |
| J | 524 | hCoV-19/England/ALDP-951DD3/2020\|EPI_ISL_551305\|20 | ···················· | ···················· | ···················· | ···················· | ···················· | ···················· |
| J | 525 | hCoV-19/England/ALDP-9520E8/2020\|EPI_ISL_551306\|20 | ···················· | ···················· | ···················· | ···················· | ···················· | ···················· |
| J | 526 | hCoV-19/England/ALDP-951F19/2020\|EPI_ISL_551307\|20 | ···················· | ···················· | ···················· | ···················· | ···················· | ···················· |
| J | 527 | hCoV-19/England/ALDP-951B33/2020\|EPI_ISL_551308\|20 | ···················· | ···················· | ···················· | ···················· | ···················· | ···················· |
| J | 528 | hCoV-19/England/ALDP-951FCE/2020\|EPI_ISL_551309\|20 | ···················· | ···················· | ···················· | ···················· | ···················· | ···················· |
| J | 529 | hCoV-19/England/ALDP-951EA3/2020\|EPI_ISL_551311\|20 | ···················· | ·A·················· | ···················· | ···················· | ···················· | ···················· |
| J | 530 | hCoV-19/England/ALDP-951F82/2020\|EPI_ISL_551313\|20 | ···················· | ···················· | ···················· | ···················· | ···················· | ···················· |
| J | 531 | hCoV-19/England/ALDP-951D97/2020\|EPI_ISL_551314\|20 | ···················· | ···················· | ···················· | ···················· | ···················· | ···················· |
| J | 532 | hCoV-19/England/ALDP-952033/2020\|EPI_ISL_551315\|20 | ···················· | ···················· | ···················· | ···················· | ···················· | ···················· |
| J | 533 | hCoV-19/England/ALDP-951BE8/2020\|EPI_ISL_551316\|20 | ···················· | ···················· | ···················· | ···················· | ···················· | ···················· |
| J | 534 | hCoV-19/England/ALDP-951CC7/2020\|EPI_ISL_551317\|20 | ···················· | ···················· | ···················· | ···················· | ···················· | ···················· |
| J | 535 | hCoV-19/England/ALDP-952112/2020\|EPI_ISL_551318\|20 | ···················· | ···················· | ···················· | ···················· | ···················· | ···················· |

| | | | | | | | | |
|---|---|---|---|---|---|---|---|---|
| J | 536 | hCoV-19/England/ALDP-952103/2020\|EPI_ISL_551319\|20 | ···················· | ···················· | ····················· | ···················· | ···················· | ···················· |
| J | 537 | hCoV-19/England/ALDP-951EB2/2020\|EPI_ISL_551320\|20 | ···················· | ·A·················· | ····················· | ···················· | ···················· | ···················· |
| J | 538 | hCoV-19/England/ALDP-951F28/2020\|EPI_ISL_551322\|20 | ···················· | ···················· | ····················· | ···················· | ···················· | ···················· |
| J | 539 | hCoV-19/England/ALDP-951C5E/2020\|EPI_ISL_551323\|20 | ···················· | ···················· | ····················· | ···················· | ···················· | ···················· |
| J | 540 | hCoV-19/England/ALDP-951E49/2020\|EPI_ISL_551324\|20 | ···················· | ···················· | ····················· | ···················· | ···················· | ···················· |
| J | 541 | hCoV-19/England/ALDP-951FA0/2020\|EPI_ISL_551325\|20 | ···················· | ···················· | ····················· | ···················· | ···················· | ···················· |
| J | 542 | hCoV-19/England/ALDP-951EEF/2020\|EPI_ISL_551326\|20 | ···················· | ···················· | ····················· | ···················· | ···················· | ···················· |
| J | 543 | hCoV-19/England/ALDP-951FFB/2020\|EPI_ISL_551328\|20 | ···················· | ·A·················· | ····················· | ···················· | ···················· | ···················· |
| J | 544 | hCoV-19/England/ALDP-951D88/2020\|EPI_ISL_551330\|20 | ···················· | ···················· | ····················· | ···················· | ···················· | ···················· |
| J | 545 | hCoV-19/England/ALDP-951C6D/2020\|EPI_ISL_551332\|20 | ···················· | ···················· | ····················· | ···················· | ···················· | ···················· |
| J | 546 | hCoV-19/England/MILK-9757F9/2020\|EPI_ISL_552610\|20 | ···················· | ···················· | ····················· | ···················· | ···················· | ···················· |
| J | 547 | hCoV-19/England/MILK-975252/2020\|EPI_ISL_552611\|20 | ···················· | ···················· | ····················· | ···················· | ···················· | ···················· |
| J | 548 | hCoV-19/England/MILK-9752CB/2020\|EPI_ISL_552612\|20 | ···················· | ···················· | ····················· | ···················· | ···················· | ···················· |
| J | 549 | hCoV-19/England/MILK-97547A/2020\|EPI_ISL_552616\|20 | ···················· | ···················· | ····················· | ···················· | ···················· | ···················· |
| J | 550 | hCoV-19/England/MILK-97546B/2020\|EPI_ISL_552621\|20 | ···················· | ···················· | ····················· | ···················· | ···················· | ···················· |
| J | 551 | hCoV-19/England/ALDP-9815F0/2020\|EPI_ISL_552622\|20 | ···················· | ···················· | ····················· | ···················· | ···················· | ···················· |
| J | 552 | hCoV-19/England/MILK-975999/2020\|EPI_ISL_552623\|20 | ···················· | ···················· | ····················· | ···················· | ···················· | ···················· |
| J | 553 | hCoV-19/England/MILK-975911/2020\|EPI_ISL_552624\|20 | ···················· | ···················· | ····················· | ···················· | ···················· | ···················· |
| J | 554 | hCoV-19/England/MILK-974AA6/2020\|EPI_ISL_552625\|20 | ···················· | ···················· | ····················· | ···················· | ···················· | ···················· |
| J | 555 | hCoV-19/England/MILK-975BA2/2020\|EPI_ISL_552626\|20 | ···················· | ·A·················· | ····················· | ···················· | ···················· | ···················· |
| J | 556 | hCoV-19/England/MILK-975841/2020\|EPI_ISL_552627\|20 | ···················· | ···················· | ····················· | ···················· | ···················· | ···················· |
| J | 557 | hCoV-19/England/MILK-97501C/2020\|EPI_ISL_552628\|20 | ···················· | ···················· | ····················· | ···················· | ···················· | ···················· |
| J | 558 | hCoV-19/England/ALDP-9829B7/2020\|EPI_ISL_552629\|20 | ···················· | ···················· | ····················· | ···················· | ···················· | ···················· |
| J | 559 | hCoV-19/Wales/MILK-975780/2020\|EPI_ISL_552630\|2020 | ···················· | ···················· | ····················· | ···················· | ···················· | ···················· |
| J | 560 | hCoV-19/England/MILK-975656/2020\|EPI_ISL_552632\|20 | ···················· | ···················· | ····················· | ···················· | ···················· | ···················· |
| J | 561 | hCoV-19/England/MILK-974FE3/2020\|EPI_ISL_552633\|20 | ···················· | ···················· | ····················· | ···················· | ···················· | ···················· |
| J | 562 | hCoV-19/England/MILK-9751EC/2020\|EPI_ISL_552634\|20 | ···················· | ···················· | ····················· | ···················· | ···················· | ···················· |
| J | 563 | hCoV-19/Wales/MILK-9750C1/2020\|EPI_ISL_552635\|2020 | ···················· | ···················· | ····················· | ···················· | ···················· | ···················· |
| J | 564 | hCoV-19/England/MILK-975BFD/2020\|EPI_ISL_552637\|20 | ···················· | ···················· | ····················· | ···················· | ···················· | ···················· |
| J | 565 | hCoV-19/England/MILK-975559/2020\|EPI_ISL_552638\|20 | ···················· | ···················· | ····················· | ···················· | ···C················ | ···················· |
| J | 566 | hCoV-19/England/ALDP-982A2D/2020\|EPI_ISL_552639\|20 | ···················· | ···················· | ····················· | ···················· | ···················· | ···················· |
| J | 567 | hCoV-19/England/MILK-9754F2/2020\|EPI_ISL_552640\|20 | ···················· | ···················· | ····················· | ···················· | ···················· | ···················· |
| J | 568 | hCoV-19/England/MILK-9750B2/2020\|EPI_ISL_552641\|20 | ···················· | ···················· | ····················· | ···················· | ···················· | ···················· |
| J | 569 | hCoV-19/England/ALDP-982832/2020\|EPI_ISL_552642\|20 | ···················· | ···················· | ····················· | ···················· | ···················· | ···················· |
| J | 570 | hCoV-19/England/MILK-974B76/2020\|EPI_ISL_552643\|20 | ···················· | ···················· | ····················· | ···················· | ···················· | ···················· |
| J | 571 | hCoV-19/England/ALDP-982AC3/2020\|EPI_ISL_552644\|20 | ···················· | ·A·················· | ····················· | ···················· | ···················· | ···················· |
| J | 572 | hCoV-19/England/ALDP-982AF0/2020\|EPI_ISL_552646\|20 | ···················· | ···················· | ····················· | ···················· | ···················· | ···················· |
| J | 573 | hCoV-19/England/MILK-97587E/2020\|EPI_ISL_552647\|20 | ···················· | ···················· | ····················· | ···················· | ···················· | ···················· |
| J | 574 | hCoV-19/England/MILK-9754C5/2020\|EPI_ISL_552650\|20 | ···················· | ···················· | ····················· | ···················· | ···················· | ···················· |
| J | 575 | hCoV-19/England/MILK-975313/2020\|EPI_ISL_552651\|20 | ···················· | ···················· | ····················· | ···················· | ···················· | ···················· |
| J | 576 | hCoV-19/England/MILK-975B93/2020\|EPI_ISL_552652\|20 | ···················· | ·A·················· | ····················· | ···················· | ···················· | ···A················ |
| J | 577 | hCoV-19/England/ALDP-982C54/2020\|EPI_ISL_552655\|20 | ···················· | ···················· | ····················· | ···················· | ···················· | ···················· |
| J | 578 | hCoV-19/England/MILK-975A3C/2020\|EPI_ISL_552656\|20 | ···················· | ···················· | ····················· | ···················· | ···················· | ···················· |
| J | 579 | hCoV-19/England/MILK-975B84/2020\|EPI_ISL_552658\|20 | ···················· | ·A·················· | ····················· | ···················· | ···················· | ···A················ |
| J | 580 | hCoV-19/England/ALDP-9828D8/2020\|EPI_ISL_552659\|20 | ···················· | ···················· | ····················· | ···················· | ···················· | ···················· |
| J | 581 | hCoV-19/England/MILK-97595D/2020\|EPI_ISL_552660\|20 | ···················· | ···················· | ····················· | ···················· | ···················· | ···················· |
| J | 582 | hCoV-19/England/MILK-975753/2020\|EPI_ISL_552661\|20 | ···················· | ···················· | ····················· | ···················· | ···················· | ···················· |
| J | 583 | hCoV-19/England/MILK-9752AD/2020\|EPI_ISL_552662\|20 | ···················· | ···················· | ····················· | ···················· | ···················· | ···················· |
| J | 584 | hCoV-19/Wales/MILK-975771/2020\|EPI_ISL_552663\|2020 | ···················· | ···················· | ····················· | ···················· | ···················· | ···················· |
| J | 585 | hCoV-19/England/ALDP-955B20/2020\|EPI_ISL_553940\|20 | ···················· | ···················· | ····················· | ···················· | ···················· | ···················· |
| J | 586 | hCoV-19/England/ALDP-955FE8/2020\|EPI_ISL_553941\|20 | ···················· | ···················· | ····················· | ···················· | ···················· | ···················· |
| J | 587 | hCoV-19/England/ALDP-956239/2020\|EPI_ISL_553942\|20 | ···················· | ···················· | ····················· | ···················· | ···················· | ···················· |
| J | 588 | hCoV-19/England/ALDP-956390/2020\|EPI_ISL_553943\|20 | ···················· | ···················· | ····················· | ···················· | ···················· | ···················· |
| J | 589 | hCoV-19/England/ALDP-95259E/2020\|EPI_ISL_553944\|20 | ···················· | ···················· | ····················· | ···················· | ···················· | ···················· |
| J | 590 | hCoV-19/England/ALDP-9565E5/2020\|EPI_ISL_553945\|20 | ···················· | ···················· | ····················· | ···················· | ···················· | ···················· |
| J | 591 | hCoV-19/England/ALDP-9527F2/2020\|EPI_ISL_553951\|20 | ···················· | ···················· | ····················· | ···················· | ···················· | ···················· |
| J | 592 | hCoV-19/England/MILK-9577DF/2020\|EPI_ISL_553952\|20 | ···················· | ···················· | ····················· | ···················· | ···················· | ···················· |
| J | 593 | hCoV-19/England/ALDP-95272F/2020\|EPI_ISL_553953\|20 | ···················· | ···················· | ····················· | ···················· | ···················· | ···················· |
| J | 594 | hCoV-19/England/MILK-957405/2020\|EPI_ISL_553954\|20 | ···················· | ···················· | ····················· | ···················· | ···················· | ···················· |
| J | 595 | hCoV-19/England/MILK-957915/2020\|EPI_ISL_553955\|20 | ···················· | ·A·················· | ····················· | ···················· | ···················· | ···················· |
| J | 596 | hCoV-19/England/MILK-95749C/2020\|EPI_ISL_553956\|20 | ···················· | ···················· | ····················· | ···················· | ···················· | ···················· |
| J | 597 | hCoV-19/England/MILK-957784/2020\|EPI_ISL_553957\|20 | ···················· | ···················· | ····················· | ···················· | ···················· | ···················· |
| J | 598 | hCoV-19/England/ALDP-9564BB/2020\|EPI_ISL_553958\|20 | ···················· | ···················· | ····················· | ···················· | ···················· | ···················· |
| J | 599 | hCoV-19/England/ALDP-952552/2020\|EPI_ISL_553959\|20 | ···················· | ···················· | ····················· | ···················· | ···················· | ···················· |
| J | 600 | hCoV-19/England/ALDP-955E45/2020\|EPI_ISL_553960\|20 | ···················· | ·A·················· | ····················· | ···················· | ···················· | ···················· |
| J | 601 | hCoV-19/England/ALDP-95269B/2020\|EPI_ISL_553962\|20 | ···················· | ···················· | ····················· | ···················· | ···················· | ···················· |
| J | 602 | hCoV-19/England/ALDP-952710/2020\|EPI_ISL_553963\|20 | ···················· | ···················· | ····················· | ···················· | ···················· | ···················· |
| J | 603 | hCoV-19/Wales/ALDP-955EEB/2020\|EPI_ISL_553964\|2020 | ···················· | ···················· | ····················· | ···················· | ···················· | ···················· |
| J | 604 | hCoV-19/England/ALDP-95658B/2020\|EPI_ISL_553966\|20 | ···················· | ···················· | ···········T········· | ···················· | ···················· | ···················· |
| J | 605 | hCoV-19/England/ALDP-955D0C/2020\|EPI_ISL_553967\|20 | ···················· | ···················· | ····················· | ···················· | ···················· | ···················· |
| J | 606 | hCoV-19/England/ALDP-9562B1/2020\|EPI_ISL_553969\|20 | ···················· | ·A·················· | ····················· | ···················· | ···················· | ···················· |
| J | 607 | hCoV-19/England/MILK-95756C/2020\|EPI_ISL_553972\|20 | ···················· | ···················· | ····················· | ···················· | ···················· | ···················· |
| J | 608 | hCoV-19/England/ALDP-955B3F/2020\|EPI_ISL_553973\|20 | ···················· | ···················· | ····················· | ···················· | ···················· | ···················· |
| J | 609 | hCoV-19/England/ALDP-9527A7/2020\|EPI_ISL_553975\|20 | ···················· | ···················· | ····················· | ···················· | ···················· | ···················· |
| J | 610 | hCoV-19/Wales/ALDP-952CD5/2020\|EPI_ISL_553976\|2020 | ···················· | ···················· | ····················· | ···················· | ···················· | ···················· |
| J | 611 | hCoV-19/England/ALDP-95273E/2020\|EPI_ISL_553977\|20 | ···················· | ···················· | ····················· | ···················· | ···················· | ···················· |
| J | 612 | hCoV-19/England/ALDP-95283B/2020\|EPI_ISL_553978\|20 | ···················· | ···················· | ····················· | ···················· | ···················· | ···················· |
| J | 613 | hCoV-19/England/ALDP-95608A/2020\|EPI_ISL_553979\|20 | ···················· | ·A·················· | ····················· | ···················· | ···················· | ···················· |
| J | 614 | hCoV-19/England/ALDP-956679/2020\|EPI_ISL_553980\|20 | ···················· | ···················· | ····················· | ···················· | ···················· | ···················· |
| J | 615 | hCoV-19/England/ALDP-952640/2020\|EPI_ISL_553981\|20 | ···················· | ···················· | ····················· | ···················· | ···················· | ···················· |

J 616 hCoV-19/England/ALDP-95666A/2020|EPI_ISL_553982|20 ···················· ···················· ····················· ···················· ···················· ····················
J 617 hCoV-19/England/MILK-9573F9/2020|EPI_ISL_553983|20 ···················· ···················· ····················· ···················· ···················· ····················
J 618 hCoV-19/England/ALDP-9529ED/2020|EPI_ISL_553984|20 ···················· ···················· ····················· ···················· ···················· ····················
J 619 hCoV-19/England/ALDP-955B02/2020|EPI_ISL_553986|20 ···················· ···················· ····················· ···················· ···················· ····················
J 620 hCoV-19/England/MILK-957C2B/2020|EPI_ISL_553987|20 ···················· ···················· ····················· ···················· ···················· ····················
J 621 hCoV-19/England/ALDP-952956/2020|EPI_ISL_553988|20 ···················· ···················· ····················· ···················· ···················· ····················
J 622 hCoV-19/England/MILK-95747E/2020|EPI_ISL_553989|20 ···················· ···················· ····················· ···················· ···················· ····················
J 623 hCoV-19/England/ALDP-956433/2020|EPI_ISL_553991|20 ···················· ···················· ····················· ···················· ···················· ····················
J 624 hCoV-19/Wales/ALDP-955ECD/2020|EPI_ISL_553992|2020 ···················· ···················· ····················· ···················· ···················· ····················
J 625 hCoV-19/England/MILK-957A30/2020|EPI_ISL_553993|20 ···················· ···················· ····················· ···················· ···················· ····················
J 626 hCoV-19/England/MILK-9576F0/2020|EPI_ISL_553994|20 ···················· ···················· ····················· ···················· ···················· ····················
J 627 hCoV-19/England/ALDP-94C9D7/2020|EPI_ISL_556002|20 ···················· ···················· ····················· ···················· ···················· ····················
J 628 hCoV-19/England/ALDP-94F4A6/2020|EPI_ISL_556003|20 ···················· ···················· ····················· ···················· ···················· ····················
J 629 hCoV-19/England/ALDP-94C597/2020|EPI_ISL_556004|20 ···················· ···················· ····················· ···················· ···················· ····················
J 630 hCoV-19/England/ALDP-94F242/2020|EPI_ISL_556006|20 ···················· ···················· ····················· ···················· ···················· ····················
J 631 hCoV-19/England/ALDP-94CF99/2020|EPI_ISL_556008|20 ···················· ···················· ····················· ···················· ···················· ····················
J 632 hCoV-19/England/ALDP-94CEC9/2020|EPI_ISL_556009|20 ···················· ···················· ····················· ···················· ···················· ····················
J 633 hCoV-19/England/ALDP-94F43D/2020|EPI_ISL_556010|20 ···················· ···················· ····················· ···················· ···················· ····················
J 634 hCoV-19/England/ALDP-94CE50/2020|EPI_ISL_556011|20 ···················· ···················· ····················· ···················· ···················· ····················
J 635 hCoV-19/England/CAMC-9499E9/2020|EPI_ISL_556012|20 ···················· ···················· ···········T········· ···················· ···················· ····················
J 636 hCoV-19/England/ALDP-94C8AD/2020|EPI_ISL_556013|20 ···················· ···················· ····················· ···················· ···················· ····················
J 637 hCoV-19/England/ALDP-94F44C/2020|EPI_ISL_556015|20 ···················· ·A·················· ····················· ···················· ···················· ····················
J 638 hCoV-19/England/ALDP-94CB3B/2020|EPI_ISL_556016|20 ···················· ···················· ····················· ···················· ···················· ····················
J 639 hCoV-19/England/ALDP-94CA2F/2020|EPI_ISL_556017|20 ···················· ···················· ····················· ···················· ···················· ····················
J 640 hCoV-19/England/ALDP-94F34F/2020|EPI_ISL_556018|20 ···················· ·············A······ ····················· ···················· ···················· ····················
J 641 hCoV-19/England/ALDP-94F51C/2020|EPI_ISL_556019|20 ···················· ···················· ····················· ···················· ···················· ····················
J 642 hCoV-19/England/ALDP-94F312/2020|EPI_ISL_556020|20 ···················· ···················· ····················· ···················· ···················· ····················
J 643 hCoV-19/England/CAMC-94ACD0/2020|EPI_ISL_556021|20 ···················· ······A············· ····················· ···················· ···················· ····················
J 644 hCoV-19/England/CAMC-948CC3/2020|EPI_ISL_556022|20 ···················· ···················· ····················· ···················· ···················· ····················
J 645 hCoV-19/England/ALDP-94C658/2020|EPI_ISL_556023|20 ···················· ···················· ····················· ···················· ···················· ····················
J 646 hCoV-19/England/ALDP-94F36D/2020|EPI_ISL_556025|20 ···················· ···················· ····················· ···················· ···················· ····················
J 647 hCoV-19/England/CAMC-94AA4F/2020|EPI_ISL_556026|20 ···················· ···················· ····················· ···················· ···················· ····················
J 648 hCoV-19/England/ALDP-94CF3F/2020|EPI_ISL_556027|20 ···················· ···················· ····················· ···················· ···················· ····················
J 649 hCoV-19/England/ALDP-94C48B/2020|EPI_ISL_556028|20 ···················· ···················· ····················· ···················· ···················· ····················
J 650 hCoV-19/England/ALDP-94C6EF/2020|EPI_ISL_556030|20 ···················· ···················· ····················· ···················· ···················· ····················
J 651 hCoV-19/England/ALDP-94F53A/2020|EPI_ISL_556031|20 ···················· ···················· ····················· ···················· ···················· ····················
J 652 hCoV-19/England/ALDP-94C4E5/2020|EPI_ISL_556032|20 ···················· ···················· ····················· ···················· ···················· ····················
J 653 hCoV-19/England/ALDP-94C579/2020|EPI_ISL_556033|20 ···················· ···················· ····················· ···················· ···················· ····················
J 654 hCoV-19/England/ALDP-94CFA8/2020|EPI_ISL_556034|20 ···················· ···················· ····················· ···················· ···················· ····················
J 655 hCoV-19/England/ALDP-94CCCF/2020|EPI_ISL_556035|20 ···················· ···················· ····················· ···················· ···················· ····················
J 656 hCoV-19/England/ALDP-94CCFC/2020|EPI_ISL_556036|20 ···················· ·A·················· ····················· ···················· ···················· ····················
J 657 hCoV-19/England/ALDP-94CCA1/2020|EPI_ISL_556037|20 ···················· ···················· ····················· ···················· ···················· ····················
J 658 hCoV-19/England/ALDP-94C9B9/2020|EPI_ISL_556038|20 ···················· ···················· ····················· ···················· ···················· ····················
J 659 hCoV-19/England/CAMC-949828/2020|EPI_ISL_556039|20 ···················· ···················· ····················· ···················· ···················· ····················
J 660 hCoV-19/England/ALDP-94C500/2020|EPI_ISL_556040|20 ···················· ·A·················· ····················· ···················· ···················· ····················
J 661 hCoV-19/England/ALDP-94CAA7/2020|EPI_ISL_556042|20 ···················· ···················· ····················· ···················· ···················· ····················
J 662 hCoV-19/England/ALDP-94C8F8/2020|EPI_ISL_556044|20 ···················· ···················· ····················· ···················· ···················· ····················
J 663 hCoV-19/England/ALDP-94F5EF/2020|EPI_ISL_556045|20 ···················· ···················· ····················· ···················· ···················· ····················
J 664 hCoV-19/England/CAMC-948E27/2020|EPI_ISL_556047|20 ···················· ···················· ····················· ···················· ···················· ····················
J 665 hCoV-19/England/ALDP-94CB1D/2020|EPI_ISL_556048|20 ···················· ···················· ····················· ···················· ···················· ····················
J 666 hCoV-19/Wales/ALDP-94CA01/2020|EPI_ISL_556049|2020 ···················· ···················· ····················· ···················· ···················· ····················
J 667 hCoV-19/England/MILK-85814A/2020|EPI_ISL_557317|20 ···················· ···················· ····················· ···················· ···················· ····················
J 668 hCoV-19/England/MILK-829D4E/2020|EPI_ISL_557318|20 ···················· ·A·················· ····················· ···················· ···················· ····················
J 669 hCoV-19/England/MILK-837EFF/2020|EPI_ISL_557319|20 ···················· ···················· ····················· ···················· ···················· ····················
J 670 hCoV-19/England/MILK-809755/2020|EPI_ISL_557321|20 ···················· ···················· ····················· ···················· ···················· ····················
J 671 hCoV-19/England/MILK-85762E/2020|EPI_ISL_557322|20 ···················· ·A·················· ····················· ···················· ···················· ···A················
J 672 hCoV-19/England/MILK-856DEE/2020|EPI_ISL_557323|20 ···················· ···················· ····················· ···················· ···················· ····················
J 673 hCoV-19/England/MILK-817C37/2020|EPI_ISL_557326|20 ···················· ···················· ····················· ···················· ···················· ····················
J 674 hCoV-19/England/MILK-82CCA8/2020|EPI_ISL_557327|20 ···················· ···················· ····················· ···················· ···················· ····················
J 675 hCoV-19/England/MILK-84D1B1/2020|EPI_ISL_557328|20 ···················· ···················· ····················· ···················· ···················· ····················
J 676 hCoV-19/England/MILK-841C9E/2020|EPI_ISL_557329|20 ···················· ···················· ····················· ···················· ···················· ····················
J 677 hCoV-19/England/MILK-856197/2020|EPI_ISL_557331|20 ···················· ···················· ····················· ···················· ···················· ····················
J 678 hCoV-19/England/MILK-84375F/2020|EPI_ISL_557332|20 ···················· ···················· ····················· ···················· ···················· ····················
J 679 hCoV-19/England/MILK-808AF3/2020|EPI_ISL_557333|20 ···················· ···················· ····················· ···················· ···················· ····················
J 680 hCoV-19/England/MILK-817A4C/2020|EPI_ISL_557334|20 ···················· ···················· ····················· ···················· ···················· ····················
J 681 hCoV-19/England/MILK-824174/2020|EPI_ISL_557336|20 ···················· ···················· ····················· ···················· ···················· ····················
J 682 hCoV-19/Wales/MILK-848FF8/2020|EPI_ISL_557337|2020 ···················· ···················· ····················· ···················· ···················· ····················
J 683 hCoV-19/England/MILK-8262F7/2020|EPI_ISL_557338|20 ···················· ···················· ····················· ···················· ···················· ····················
J 684 hCoV-19/England/MILK-813E9F/2020|EPI_ISL_557339|20 ···················· ···················· ····················· ···················· ···················· ····················
J 685 hCoV-19/England/MILK-82B571/2020|EPI_ISL_557341|20 ···················· ···················· ····················· ···················· ···················· ····················
J 686 hCoV-19/England/MILK-878F95/2020|EPI_ISL_557342|20 ···················· ···················· ····················· ···················· ···················· ····················
J 687 hCoV-19/England/MILK-83B7B4/2020|EPI_ISL_557343|20 ···················· ···················· ····················· ···················· ···················· ····················
J 688 hCoV-19/England/MILK-8327EA/2020|EPI_ISL_557344|20 ···················· ···················· ····················· ···················· ···················· ····················
J 689 hCoV-19/England/MILK-80C2F7/2020|EPI_ISL_557346|20 ···················· ·············A······ ····················· ···················· ···················· ····················
J 690 hCoV-19/England/MILK-83D8EC/2020|EPI_ISL_557347|20 ···················· ···················· ····················· ···················· ···················· ····················
J 691 hCoV-19/England/MILK-828608/2020|EPI_ISL_557348|20 ···················· ···················· ····················· ···················· ···················· ····················
J 692 hCoV-19/England/MILK-8524A1/2020|EPI_ISL_557349|20 ···················· ···················· ····················· ···················· ···················· ····················
J 693 hCoV-19/England/MILK-875DCB/2020|EPI_ISL_557350|20 ···················· ···················· ····················· ···················· ···················· ····················
J 694 hCoV-19/Wales/ALDP-6FA9F9/2020|EPI_ISL_557351|2020 ···················· ···················· ····················· ···················· ···················· ····················
J 695 hCoV-19/England/MILK-852A09/2020|EPI_ISL_557352|20 ···················· ···················· ····················· ···················· ···················· ····················

J 696 hCoV-19/England/ALDP-745C22/2020|EPI_ISL_557354|20 ···················· ···················· ···················· ···················· ···················· ····················
J 697 hCoV-19/England/ALDP-6FB8FB/2020|EPI_ISL_557356|20 ···················· ·A·················· ···················· ···················· ···················· ····················
J 698 hCoV-19/England/MILK-6F2D8C/2020|EPI_ISL_557357|20 ···················· ·A·················· ···················· ···················· ···················· ····················
J 699 hCoV-19/England/ALDP-744B9F/2020|EPI_ISL_557359|20 ···················· ···················· ···················· ···················· ···················· ····················
J 700 hCoV-19/England/MILK-85C1DD/2020|EPI_ISL_557360|20 ···················· ···················· ···················· ···················· ···················· ····················
J 701 hCoV-19/England/ALDP-74622F/2020|EPI_ISL_557361|20 ···················· ···················· ···················· ···················· ···················· ····················
J 702 hCoV-19/England/ALDP-6FC9D9/2020|EPI_ISL_557363|20 ···················· ···················· ···················· ···················· ···················· ····················
J 703 hCoV-19/England/MILK-85FE79/2020|EPI_ISL_557364|20 ···················· ···················· ···················· ···················· ···················· ····················
J 704 hCoV-19/England/MILK-86F043/2020|EPI_ISL_557365|20 ···················· ·A·················· ···················· ···················· ···················· ····················
J 705 hCoV-19/England/MILK-74B0F3/2020|EPI_ISL_557367|20 ···················· ···················· ···················· ···················· ···················· ····················
J 706 hCoV-19/England/MILK-6F7B8D/2020|EPI_ISL_557369|20 ···················· ···················· ···················· ···················· ···················· ····················
J 707 hCoV-19/England/MILK-85D321/2020|EPI_ISL_557372|20 ···················· ···················· ···················· ···················· ···················· ····················
J 708 hCoV-19/England/ALDP-6FBFF6/2020|EPI_ISL_557373|20 ···················· ···················· ···················· ···················· ···················· ····················
J 709 hCoV-19/England/ALDP-744DB7/2020|EPI_ISL_557374|20 ···················· ···················· ···················· ···················· ···················· ····················
J 710 hCoV-19/England/MILK-74CE52/2020|EPI_ISL_557375|20 ···················· ···················· ···················· ···················· ···················· ····················
J 711 hCoV-19/England/MILK-876BA2/2020|EPI_ISL_557377|20 ···················· ···················· ···················· ···················· ···················· ····················
J 712 hCoV-19/England/ALDP-6FE6D0/2020|EPI_ISL_557378|20 ···················· ···················· ···················· ···················· ···················· ····················
J 713 hCoV-19/England/ALDP-5AF0FF/2020|EPI_ISL_558648|20 ···················· ···················· ···················· ···················· ···················· ····················
J 714 hCoV-19/England/ALDP-5B5E38/2020|EPI_ISL_558649|20 ···················· ···················· ···················· ···················· ···················· ····················
J 715 hCoV-19/England/ALDP-5A6A63/2020|EPI_ISL_558650|20 ···················· ···················· ···················· ···················· ···················· ····················
J 716 hCoV-19/England/ALDP-5B0240/2020|EPI_ISL_558651|20 ···················· ···················· ···················· ···················· ···················· ····················
J 717 hCoV-19/England/ALDP-5ACDA0/2020|EPI_ISL_558652|20 ···················· ···················· ···················· ···················· ···················· ····················
J 718 hCoV-19/England/ALDP-5A3B63/2020|EPI_ISL_558654|20 ···················· ···················· ···················· ···················· ···················· ····················
J 719 hCoV-19/England/ALDP-5C2D1D/2020|EPI_ISL_558655|20 ···················· ···················· ···················· ···················· ···················· ····················
J 720 hCoV-19/England/ALDP-5A97A5/2020|EPI_ISL_558656|20 ···················· ···················· ···················· ···················· ···················· ····················
J 721 hCoV-19/England/ALDP-5AA908/2020|EPI_ISL_558657|20 ···················· ···················· ···················· ···················· ···················· ····················
J 722 hCoV-19/England/ALDP-5AB257/2020|EPI_ISL_558658|20 ···················· ···················· ···················· ···················· ···················· ····················
J 723 hCoV-19/England/ALDP-5B76C5/2020|EPI_ISL_558659|20 ···················· ···················· ···················· ···················· ···················· ····················
J 724 hCoV-19/England/ALDP-5A41D9/2020|EPI_ISL_558660|20 ···················· ···················· ···················· ···················· ···················· ····················
J 725 hCoV-19/England/ALDP-59EB4D/2020|EPI_ISL_558662|20 ···················· ···················· ···················· ···················· ···················· ····················
J 726 hCoV-19/England/ALDP-5B49DE/2020|EPI_ISL_558663|20 ···················· ···················· ···················· ···················· ···················· ····················
J 727 hCoV-19/England/ALDP-5BE172/2020|EPI_ISL_558664|20 ···················· ···················· ···················· ···················· ···················· ····················
J 728 hCoV-19/England/ALDP-59F827/2020|EPI_ISL_558665|20 ···················· ···················· ···················· ···················· ···················· ····················
J 729 hCoV-19/England/ALDP-5A7868/2020|EPI_ISL_558668|20 ···················· ···················· ···················· ···················· ···················· ····················
J 730 hCoV-19/England/ALDP-5A56CA/2020|EPI_ISL_558669|20 ···················· ···················· ···················· ···················· ···················· ····················
J 731 hCoV-19/England/ALDP-5ADE51/2020|EPI_ISL_558670|20 ···················· ···················· ···················· ···················· ···················· ····················
J 732 hCoV-19/England/ALDP-5A329B/2020|EPI_ISL_558671|20 ···················· ···················· ···················· ···················· ···················· ····················
J 733 hCoV-19/England/ALDP-5BDE7C/2020|EPI_ISL_558672|20 ···················· ·············A······ ···················· ···················· ···················· ····················
J 734 hCoV-19/England/ALDP-5B475C/2020|EPI_ISL_558673|20 ···················· ···················· ···················· ···················· ···················· ····················
J 735 hCoV-19/England/ALDP-5A0DCA/2020|EPI_ISL_558674|20 ···················· ···················· ···················· ···················· ···················· ····················
J 736 hCoV-19/England/ALDP-5A5563/2020|EPI_ISL_558675|20 ···················· ···················· ···················· ···················· ···················· ····················
J 737 hCoV-19/England/ALDP-5AB9E9/2020|EPI_ISL_558676|20 ···················· ···················· ···················· ···················· ···················· ····················
J 738 hCoV-19/England/ALDP-5AA971/2020|EPI_ISL_558677|20 ···················· ···················· ···················· ···················· ···················· ····················
J 739 hCoV-19/England/ALDP-5A67E4/2020|EPI_ISL_558678|20 ···················· ···················· ···················· ···················· ···················· ····················
J 740 hCoV-19/England/ALDP-5A9F9E/2020|EPI_ISL_558680|20 ···················· ···················· ···················· ···················· ···················· ····················
J 741 hCoV-19/Wales/ALDP-5A2EB6/2020|EPI_ISL_558681|2020 ···················· ···················· ···················· ···················· ···················· ····················
J 742 hCoV-19/England/ALDP-5A1F69/2020|EPI_ISL_558682|20 ···················· ·A·················· ···················· ···················· ···················· ····················
J 743 hCoV-19/England/ALDP-59DDFD/2020|EPI_ISL_558683|20 ···················· ···················· ···················· ···················· ···················· ····················
J 744 hCoV-19/England/ALDP-59EA40/2020|EPI_ISL_558685|20 ···················· ···················· ···················· ···················· ···················· ····················
J 745 hCoV-19/England/ALDP-5A376F/2020|EPI_ISL_558686|20 ···················· ···················· ···················· ···················· ···················· ····················
J 746 hCoV-19/England/ALDP-5C41E2/2020|EPI_ISL_558687|20 ···················· ···················· ···················· ···················· ···················· ····················
J 747 hCoV-19/England/ALDP-59DE27/2020|EPI_ISL_558688|20 ···················· ···················· ···················· ···················· ···················· ····················
J 748 hCoV-19/England/ALDP-5AAF06/2020|EPI_ISL_558689|20 ···················· ···················· ···················· ···················· ···················· ····················
J 749 hCoV-19/England/ALDP-5A94F9/2020|EPI_ISL_558690|20 ···················· ···················· ···················· ···················· ···················· ····················
J 750 hCoV-19/England/ALDP-5B6F8F/2020|EPI_ISL_558691|20 ···················· ···················· ···················· ···················· ···················· ····················
J 751 hCoV-19/Scotland/EDB7763/2020|EPI_ISL_559979|2020- ···················· ···················· ···················· ···················· ···················· ····················
J 752 hCoV-19/Scotland/EDB7765/2020|EPI_ISL_559980|2020- ···················· ···················· ···················· ···················· ···················· ····················
J 753 hCoV-19/Scotland/EDB7768/2020|EPI_ISL_559981|2020- ···················· ···················· ···················· ···················· ·····T·············· ····················
J 754 hCoV-19/Scotland/EDB7769/2020|EPI_ISL_559982|2020- ···················· ·A·················· ···················· ···················· ···················· ····················
J 755 hCoV-19/Scotland/QEUH-996798/2020|EPI_ISL_559984|2 ···················· ·A·················· ···················· ···················· ···················· ··A·················
J 756 hCoV-19/Scotland/QEUH-9967F2/2020|EPI_ISL_559985|2 ···················· ···················· ···················· ···················· ···················· ····················
J 757 hCoV-19/Scotland/QEUH-9968C2/2020|EPI_ISL_559986|2 ···················· ·A·················· ···················· ···················· ···················· ··A·················
J 758 hCoV-19/Scotland/QEUH-9968D1/2020|EPI_ISL_559987|2 ···················· ···················· ···················· ···················· ···················· ····················
J 759 hCoV-19/Scotland/QEUH-9968FF/2020|EPI_ISL_559989|2 ···················· ···················· ···················· ···················· ···················· ····················
J 760 hCoV-19/Scotland/QEUH-996929/2020|EPI_ISL_559990|2 ···················· ···················· ···················· ···················· ···················· ····················
J 761 hCoV-19/Scotland/QEUH-997A9E/2020|EPI_ISL_559991|2 ···················· ···················· ···················· ···················· ···················· ····················
J 762 hCoV-19/Scotland/QEUH-997AAD/2020|EPI_ISL_559992|2 ···················· ···················· ···················· ···················· ···················· ····················
J 763 hCoV-19/Scotland/QEUH-997BC8/2020|EPI_ISL_559993|2 ···················· ···················· ···················· ···················· ···················· ····················
J 764 hCoV-19/Scotland/QEUH-997BE6/2020|EPI_ISL_559994|2 ···················· ···················· ···················· ···················· ···················· ····················
J 765 hCoV-19/Scotland/QEUH-99B4C9/2020|EPI_ISL_559995|2 ···················· ···················· ···················· ···················· ···················· ····················
J 766 hCoV-19/Scotland/QEUH-99B4D8/2020|EPI_ISL_559996|2 ···················· ···················· ···················· ···················· ···················· ····················
J 767 hCoV-19/Scotland/QEUH-99B62D/2020|EPI_ISL_559997|2 ···················· ···················· ···················· ···················· ···················· ····················
J 768 hCoV-19/Scotland/QEUH-99BE07/2020|EPI_ISL_559999|2 ···················· ···················· ···················· ···················· ···················· ····················
J 769 hCoV-19/England/MILK-9AB4F3/2020|EPI_ISL_551179|20 ···················· ·A·················· ···················· ···················· ···················· ····················
J 770 hCoV-19/England/MILK-9A9A11/2020|EPI_ISL_551180|20 ···················· ···················· ···················· ···················· ···················· ····················
J 771 hCoV-19/England/MILK-9A8F22/2020|EPI_ISL_551182|20 ···················· ···················· ···················· ···················· ···················· ····················
J 772 hCoV-19/England/MILK-9A9D27/2020|EPI_ISL_551183|20 ···················· ···················· ···················· ···················· ···················· ····················
J 773 hCoV-19/England/MILK-9A9CDF/2020|EPI_ISL_551184|20 ···················· ···················· ···················· ···················· ···················· ····················
J 774 hCoV-19/England/MILK-9A93F8/2020|EPI_ISL_551185|20 ···················· ···················· ···················· ···················· ···················· ····················
J 775 hCoV-19/England/MILK-9A91C1/2020|EPI_ISL_551186|20 ···················· ···················· ···················· ···················· ···················· ····················

| | | | | | | | | |
|---|---|---|---|---|---|---|---|---|
| J | 776 | hCoV-19/England/MILK-9A94E6/2020\|EPI_ISL_551187\|20 | ···················· | ·A·················· | ····················· | ···················· | ···················· | ···A················ |
| J | 777 | hCoV-19/England/MILK-9A9282/2020\|EPI_ISL_551188\|20 | ···················· | ···················· | ····················· | ···················· | ···················· | ···················· |
| J | 778 | hCoV-19/England/MILK-9A9D63/2020\|EPI_ISL_551189\|20 | ···················· | ···················· | ····················· | ···················· | ···················· | ···················· |
| J | 779 | hCoV-19/England/MILK-9A9228/2020\|EPI_ISL_551190\|20 | ···················· | ···················· | ····················· | ···················· | ···················· | ···················· |
| J | 780 | hCoV-19/England/MILK-9A9668/2020\|EPI_ISL_551192\|20 | ···················· | ···················· | ····················· | ···················· | ···················· | ···················· |
| J | 781 | hCoV-19/England/MILK-9A9677/2020\|EPI_ISL_551193\|20 | ···················· | ···················· | ····················· | ···················· | ···················· | ···················· |
| J | 782 | hCoV-19/England/MILK-9AB129/2020\|EPI_ISL_551194\|20 | ···················· | ···················· | ····················· | ···················· | ···················· | ···················· |
| J | 783 | hCoV-19/England/MILK-9A9E06/2020\|EPI_ISL_551196\|20 | ···················· | ···················· | ····················· | ···················· | ···················· | ···················· |
| J | 784 | hCoV-19/England/MILK-9AB138/2020\|EPI_ISL_551197\|20 | ···················· | ·A·················· | ····················· | ···················· | ···················· | ···A················ |
| J | 785 | hCoV-19/England/MILK-9AB46C/2020\|EPI_ISL_551198\|20 | ···················· | ·A·················· | ····················· | ···················· | ···················· | ···················· |
| J | 786 | hCoV-19/England/MILK-9AB262/2020\|EPI_ISL_551199\|20 | ···················· | ···················· | ····················· | ···················· | ···················· | ···················· |
| J | 787 | hCoV-19/England/MILK-9A9B5A/2020\|EPI_ISL_551212\|20 | ···················· | ···················· | ····················· | ···················· | ···················· | ···················· |
| J | 788 | hCoV-19/England/MILK-9A91A3/2020\|EPI_ISL_551213\|20 | ···················· | ···················· | ····················· | ···················· | ···················· | ···················· |
| J | 789 | hCoV-19/England/MILK-9A9A7B/2020\|EPI_ISL_551214\|20 | ···················· | ···················· | ····················· | ···················· | ···················· | ···················· |
| J | 790 | hCoV-19/England/MILK-9A8E25/2020\|EPI_ISL_551216\|20 | ···················· | ···················· | ····················· | ···················· | ···················· | ···················· |
| J | 791 | hCoV-19/England/MILK-9AB0FF/2020\|EPI_ISL_551217\|20 | ···················· | ···················· | ····················· | ···················· | ···················· | ···················· |
| J | 792 | hCoV-19/England/MILK-9A91D0/2020\|EPI_ISL_551220\|20 | ···················· | ···················· | ····················· | ···················· | ···················· | ···················· |
| J | 793 | hCoV-19/England/MILK-9AAE9C/2020\|EPI_ISL_551221\|20 | ········T··········· | ···················· | ····················· | ···················· | ···················· | ···················· |
| J | 794 | hCoV-19/England/MILK-9A904C/2020\|EPI_ISL_551222\|20 | ···················· | ···················· | ····················· | ···················· | ···················· | ···················· |
| J | 795 | hCoV-19/England/MILK-9A9792/2020\|EPI_ISL_551223\|20 | ···················· | ···················· | ····················· | ···················· | ···················· | ···················· |
| J | 796 | hCoV-19/England/MILK-9A97ED/2020\|EPI_ISL_551224\|20 | ···················· | ·A·················· | ····················· | ···················· | ···················· | ···················· |
| J | 797 | hCoV-19/England/MILK-9A9AD5/2020\|EPI_ISL_551225\|20 | ···················· | ···················· | ····················· | ···················· | ···················· | ···················· |
| J | 798 | hCoV-19/England/MILK-9AB52D/2020\|EPI_ISL_551227\|20 | ···················· | ···················· | ····················· | ···················· | ···················· | ···················· |
| J | 799 | hCoV-19/England/MILK-9A9501/2020\|EPI_ISL_551228\|20 | ···················· | ···················· | ····················· | ···················· | ···················· | ···················· |
| J | 800 | hCoV-19/England/MILK-9AB45D/2020\|EPI_ISL_551229\|20 | ···················· | ···················· | ····················· | ···················· | ···················· | ···················· |
| J | 801 | hCoV-19/England/MILK-9AAF8A/2020\|EPI_ISL_551230\|20 | ···················· | ···················· | ····················· | ···················· | ···················· | ···················· |
| J | 802 | hCoV-19/England/MILK-9A964A/2020\|EPI_ISL_551231\|20 | ···················· | ···················· | ····················· | ···················· | ···················· | ···················· |
| J | 803 | hCoV-19/England/MILK-9A9B3C/2020\|EPI_ISL_551232\|20 | ···················· | ···················· | ····················· | ···················· | ···················· | ···················· |
| J | 804 | hCoV-19/England/MILK-9A9738/2020\|EPI_ISL_551233\|20 | ···················· | ···················· | ····················· | ···················· | ···················· | ···················· |
| J | 805 | hCoV-19/England/MILK-9A8F40/2020\|EPI_ISL_551234\|20 | ···················· | ···················· | ····················· | ···················· | ···················· | ···················· |
| J | 806 | hCoV-19/England/MILK-9A9B2D/2020\|EPI_ISL_551236\|20 | ···················· | ···················· | ····················· | ···················· | ···················· | ···················· |
| J | 807 | hCoV-19/England/MILK-9AB235/2020\|EPI_ISL_551237\|20 | ···················· | ···················· | ····················· | ···················· | ···················· | ···················· |
| J | 808 | hCoV-19/England/MILK-9A955C/2020\|EPI_ISL_551238\|20 | ···················· | ···················· | ····················· | ···················· | ···················· | ···················· |
| J | 809 | hCoV-19/England/MILK-9A95E3/2020\|EPI_ISL_551239\|20 | ···················· | ···················· | ····················· | ···················· | ······T············· | ···················· |
| J | 810 | hCoV-19/England/MILK-9A9E60/2020\|EPI_ISL_551240\|20 | ···················· | ···················· | ····················· | ···················· | ···················· | ···················· |
| J | 811 | hCoV-19/England/MILK-9AB03B/2020\|EPI_ISL_551241\|20 | ···················· | ···················· | ····················· | ···················· | ···················· | ···················· |
| J | 812 | hCoV-19/England/MILK-9A9C66/2020\|EPI_ISL_551242\|20 | ···················· | ···················· | ····················· | ···················· | ···················· | ···················· |
| J | 813 | hCoV-19/England/MILK-9A9D81/2020\|EPI_ISL_551243\|20 | ···················· | ···················· | ····················· | ···················· | ···················· | ···················· |
| J | 814 | hCoV-19/Wales/MILK-9A9C1B/2020\|EPI_ISL_551244\|2020 | ···················· | ···················· | ····················· | ···················· | ···················· | ···················· |
| J | 815 | hCoV-19/England/MILK-9AB29F/2020\|EPI_ISL_551245\|20 | ···················· | ···················· | ····················· | ···················· | ···················· | ···················· |
| J | 816 | hCoV-19/England/MILK-9A9695/2020\|EPI_ISL_551246\|20 | ···················· | ···················· | ····················· | ···················· | ···················· | ···················· |
| J | 817 | hCoV-19/England/MILK-9A9404/2020\|EPI_ISL_551247\|20 | ···················· | ···················· | ····················· | ···················· | ···················· | ···················· |
| J | 818 | hCoV-19/England/MILK-9A99BA/2020\|EPI_ISL_551248\|20 | ···················· | ·A·················· | ····················· | ···················· | ···················· | ···················· |
| J | 819 | hCoV-19/England/MILK-9A952F/2020\|EPI_ISL_551250\|20 | ···················· | ····A··············· | ····················· | ···················· | ···················· | ···················· |
| J | 820 | hCoV-19/England/MILK-9A90C4/2020\|EPI_ISL_551251\|20 | ···················· | ···················· | ····················· | ···················· | ···················· | ···················· |
| J | 821 | hCoV-19/England/MILK-9A96FF/2020\|EPI_ISL_551252\|20 | ···················· | ···················· | ····················· | ···················· | ···················· | ···················· |
| J | 822 | hCoV-19/England/MILK-9AB47B/2020\|EPI_ISL_551253\|20 | ···················· | ···················· | ····················· | ···················· | ···················· | ···················· |
| J | 823 | hCoV-19/England/MILK-9AB305/2020\|EPI_ISL_551255\|20 | ···················· | ···················· | ····················· | ···················· | ···················· | ···················· |
| J | 824 | hCoV-19/England/MILK-9A97A1/2020\|EPI_ISL_551257\|20 | ···················· | ···················· | ····················· | ···················· | ···················· | ···················· |
| J | 825 | hCoV-19/England/MILK-9AB4A8/2020\|EPI_ISL_551260\|20 | ···················· | ···················· | ····················· | ···················· | ···················· | ···················· |
| J | 826 | hCoV-19/England/MILK-9A9686/2020\|EPI_ISL_551261\|20 | ···················· | ···················· | ····················· | ···················· | ···················· | ···················· |
| J | 827 | hCoV-19/England/MILK-9A9774/2020\|EPI_ISL_551262\|20 | ···················· | ···················· | ····················· | ···················· | ···················· | ···················· |
| J | 828 | hCoV-19/Wales/MILK-9A8E43/2020\|EPI_ISL_551263\|2020 | ···················· | ···················· | ····················· | ···················· | ···················· | ···················· |
| J | 829 | hCoV-19/England/MILK-9A902E/2020\|EPI_ISL_551264\|20 | ···················· | ·A·················· | ····················· | ···················· | ···················· | ···················· |
| J | 830 | hCoV-19/England/MILK-9A9D18/2020\|EPI_ISL_551265\|20 | ···················· | ···················· | ····················· | ···················· | ···················· | ···················· |
| J | 831 | hCoV-19/England/MILK-9A948C/2020\|EPI_ISL_551266\|20 | ···················· | ···················· | ····················· | ···················· | ···················· | ···················· |
| J | 832 | hCoV-19/England/MILK-97537D/2020\|EPI_ISL_552544\|20 | ···················· | ···················· | ····················· | ···················· | ···················· | ···················· |
| J | 833 | hCoV-19/England/MILK-975577/2020\|EPI_ISL_552545\|20 | ···················· | ···················· | ····················· | ···················· | ···················· | ···················· |
| J | 834 | hCoV-19/England/MILK-974DF8/2020\|EPI_ISL_552548\|20 | ···················· | ·A·················· | ····················· | ···················· | ···················· | ···A················ |
| J | 835 | hCoV-19/England/MILK-974B3A/2020\|EPI_ISL_552549\|20 | ···················· | ···················· | ····················· | ···················· | ···················· | ···················· |
| J | 836 | hCoV-19/England/MILK-9759D5/2020\|EPI_ISL_552550\|20 | ···················· | ···················· | ····················· | ···················· | ···················· | ···················· |
| J | 837 | hCoV-19/England/MILK-975AC3/2020\|EPI_ISL_552551\|20 | ···················· | ···················· | ····················· | ···················· | ···················· | ···················· |
| J | 838 | hCoV-19/England/MILK-97588D/2020\|EPI_ISL_552553\|20 | ···················· | ···················· | ····················· | ···················· | ···················· | ···················· |
| J | 839 | hCoV-19/England/MILK-9759A8/2020\|EPI_ISL_552554\|20 | ···················· | ·A·················· | ····················· | ···················· | ···················· | ···················· |
| J | 840 | hCoV-19/England/MILK-975AF0/2020\|EPI_ISL_552555\|20 | ···················· | ···················· | ····················· | ···················· | ···················· | ···················· |
| J | 841 | hCoV-19/England/ALDP-982850/2020\|EPI_ISL_552556\|20 | ···················· | ···················· | ····················· | ···················· | ···················· | ···················· |
| J | 842 | hCoV-19/England/MILK-97529E/2020\|EPI_ISL_552557\|20 | ···················· | ···················· | ····················· | ···················· | ···················· | ···················· |
| J | 843 | hCoV-19/England/MILK-97528F/2020\|EPI_ISL_552558\|20 | ···················· | ···················· | ····················· | ···················· | ···················· | ···················· |
| J | 844 | hCoV-19/England/MILK-97551D/2020\|EPI_ISL_552561\|20 | ···················· | ···················· | ····················· | ···················· | ···C················ | ···················· |
| J | 845 | hCoV-19/England/ALDP-982AA5/2020\|EPI_ISL_552562\|20 | ···················· | ···················· | ····················· | ···················· | ···················· | ···················· |
| J | 846 | hCoV-19/England/MILK-975173/2020\|EPI_ISL_552563\|20 | ···················· | ···················· | ····················· | ···················· | ···················· | ···················· |
| J | 847 | hCoV-19/England/ALDP-982A0F/2020\|EPI_ISL_552565\|20 | ···················· | ···················· | ····················· | ···················· | ···················· | ···················· |
| J | 848 | hCoV-19/England/ALDP-9818CA/2020\|EPI_ISL_552566\|20 | ···················· | ···················· | ····················· | ···················· | ···················· | ···················· |
| J | 849 | hCoV-19/England/MILK-9750EF/2020\|EPI_ISL_552567\|20 | ···················· | ···················· | ····················· | ···················· | ···················· | ···················· |
| J | 850 | hCoV-19/England/MILK-975322/2020\|EPI_ISL_552568\|20 | ···················· | ···················· | ····················· | ···················· | ···················· | ···················· |
| J | 851 | hCoV-19/England/MILK-974FD4/2020\|EPI_ISL_552569\|20 | ···················· | ···················· | ····················· | ···················· | ···················· | ···················· |
| J | 852 | hCoV-19/England/MILK-97503A/2020\|EPI_ISL_552571\|20 | ···················· | ···················· | ····················· | ···················· | ···················· | ···················· |
| J | 853 | hCoV-19/England/ALDP-982CDC/2020\|EPI_ISL_552574\|20 | ···················· | ···················· | ····················· | ···················· | ···················· | ···················· |
| J | 854 | hCoV-19/England/ALDP-9814B7/2020\|EPI_ISL_552577\|20 | ···················· | ···················· | ····················· | ···················· | ···················· | ···················· |
| J | 855 | hCoV-19/England/ALDP-982B75/2020\|EPI_ISL_552579\|20 | ···················· | ···················· | ····················· | ···················· | ···················· | ···················· |

| | | | | | | | |
|---|---|---|---|---|---|---|---|
| J | 856 hCoV-19/England/ALDP-982CFA/2020\|EPI_ISL_552580\|20 | ···················· | ···················· | ····················· | ···················· | ···················· | ···················· |
| J | 857 hCoV-19/England/MILK-974AD3/2020\|EPI_ISL_552581\|20 | ···················· | ···················· | ····················· | ···················· | ···················· | ···················· |
| J | 858 hCoV-19/England/MILK-97554A/2020\|EPI_ISL_552582\|20 | ···················· | ···················· | ····················· | ···················· | ···················· | ···················· |
| J | 859 hCoV-19/England/ALDP-982D15/2020\|EPI_ISL_552583\|20 | ···················· | ···················· | ····················· | ···················· | ···················· | ···················· |
| J | 860 hCoV-19/England/MILK-975498/2020\|EPI_ISL_552584\|20 | ···················· | ···················· | ····················· | ···················· | ···················· | ···················· |
| J | 861 hCoV-19/England/MILK-975234/2020\|EPI_ISL_552585\|20 | ···················· | ···················· | ····················· | ···················· | ···················· | ···················· |
| J | 862 hCoV-19/England/MILK-974EE6/2020\|EPI_ISL_552586\|20 | ···················· | ···················· | ····················· | ···················· | ···················· | ···················· |
| J | 863 hCoV-19/England/ALDP-982A87/2020\|EPI_ISL_552590\|20 | ···················· | ···················· | ····················· | ···················· | ···················· | ···················· |
| J | 864 hCoV-19/England/ALDP-98288D/2020\|EPI_ISL_552591\|20 | ···················· | ···················· | ····················· | ···················· | ···················· | ···················· |
| J | 865 hCoV-19/England/MILK-975BDF/2020\|EPI_ISL_552594\|20 | ···················· | ···················· | ····················· | ···················· | ···················· | ···················· |
| J | 866 hCoV-19/England/ALDP-9829A8/2020\|EPI_ISL_552595\|20 | ···················· | ···················· | ····················· | ···················· | ···················· | ···················· |
| J | 867 hCoV-19/England/ALDP-98154B/2020\|EPI_ISL_552596\|20 | ···················· | ···················· | ····················· | ···················· | ···················· | ···················· |
| J | 868 hCoV-19/England/MILK-975692/2020\|EPI_ISL_552597\|20 | ···················· | ···················· | ····················· | ···················· | ···················· | ···················· |
| J | 869 hCoV-19/England/ALDP-9563EB/2020\|EPI_ISL_553875\|20 | ···················· | ···················· | ····················· | ···················· | ···················· | ···················· |
| J | 870 hCoV-19/England/ALDP-955AE7/2020\|EPI_ISL_553876\|20 | ···················· | ···················· | ····················· | ···················· | ···················· | ···················· |
| J | 871 hCoV-19/England/ALDP-952BF6/2020\|EPI_ISL_553877\|20 | ···················· | ···················· | ····················· | ···················· | ···················· | ···················· |
| J | 872 hCoV-19/England/ALDP-955AC9/2020\|EPI_ISL_553878\|20 | ···················· | ···················· | ····················· | ···················· | ···················· | ···················· |
| J | 873 hCoV-19/England/MILK-9578DC/2020\|EPI_ISL_553879\|20 | ···················· | ·A·················· | ····················· | ···················· | ···················· | ···················· |
| J | 874 hCoV-19/England/MILK-95771B/2020\|EPI_ISL_553880\|20 | ···················· | ···················· | ····················· | ···················· | ···················· | ···················· |
| J | 875 hCoV-19/England/ALDP-956530/2020\|EPI_ISL_553881\|20 | ···················· | ···················· | ····················· | ···················· | ···················· | ···················· |
| J | 876 hCoV-19/England/ALDP-9526AA/2020\|EPI_ISL_553882\|20 | ···················· | ···················· | ····················· | ···················· | ···················· | ···················· |
| J | 877 hCoV-19/England/ALDP-9529CF/2020\|EPI_ISL_553883\|20 | ···················· | ···················· | ····················· | ···················· | ···················· | ···················· |
| J | 878 hCoV-19/England/ALDP-956275/2020\|EPI_ISL_553884\|20 | ···················· | ···················· | ····················· | ···················· | ···················· | ···················· |
| J | 879 hCoV-19/England/MILK-957450/2020\|EPI_ISL_553885\|20 | ···················· | ···················· | ····················· | ···················· | ···················· | ···················· |
| J | 880 hCoV-19/England/ALDP-956327/2020\|EPI_ISL_553887\|20 | ···················· | ···················· | ····················· | ···················· | ···················· | ···················· |
| J | 881 hCoV-19/England/ALDP-952AAE/2020\|EPI_ISL_553888\|20 | ···················· | ···················· | ····················· | ···················· | ···················· | ···················· |
| J | 882 hCoV-19/England/MILK-957502/2020\|EPI_ISL_553889\|20 | ···················· | ···················· | ····················· | ···················· | ···················· | ···················· |
| J | 883 hCoV-19/England/ALDP-95663D/2020\|EPI_ISL_553890\|20 | ···················· | ···················· | ····················· | ···················· | ···················· | ···················· |
| J | 884 hCoV-19/England/ALDP-955D39/2020\|EPI_ISL_553891\|20 | ···················· | ···················· | ····················· | ···················· | ···················· | ···················· |
| J | 885 hCoV-19/England/ALDP-9560E4/2020\|EPI_ISL_553892\|20 | ···················· | ···················· | ····················· | ···················· | ···················· | ···················· |
| J | 886 hCoV-19/England/MILK-957872/2020\|EPI_ISL_553893\|20 | ···················· | ·A·················· | ····················· | ···················· | ···················· | ···················· |
| J | 887 hCoV-19/England/MILK-957809/2020\|EPI_ISL_553894\|20 | ···················· | ···················· | ····················· | ···················· | ···················· | ···················· |
| J | 888 hCoV-19/England/ALDP-9527B6/2020\|EPI_ISL_553895\|20 | ···················· | ···················· | ····················· | ···················· | ···················· | ···················· |
| J | 889 hCoV-19/England/MILK-9578FA/2020\|EPI_ISL_553896\|20 | ···················· | ···················· | ····················· | ···················· | ···················· | ···················· |
| J | 890 hCoV-19/England/MILK-957854/2020\|EPI_ISL_553897\|20 | ···················· | ···················· | ····················· | ···················· | ···················· | ···················· |
| J | 891 hCoV-19/England/MILK-9573AE/2020\|EPI_ISL_553898\|20 | ···················· | ···················· | ····················· | ···················· | ···················· | ···················· |
| J | 892 hCoV-19/England/ALDP-956688/2020\|EPI_ISL_553899\|20 | ···················· | ···················· | ····················· | ···················· | ···················· | ···················· |
| J | 893 hCoV-19/England/ALDP-94F6CE/2020\|EPI_ISL_556050\|20 | ···················· | ···················· | ····················· | ···················· | ···················· | ···················· |
| J | 894 hCoV-19/England/ALDP-94CFF3/2020\|EPI_ISL_556051\|20 | ···················· | ···················· | ····················· | ···················· | ···················· | ···················· |
| J | 895 hCoV-19/England/ALDP-94CF7B/2020\|EPI_ISL_556052\|20 | ···················· | ···················· | ····················· | ···················· | ···················· | ···················· |
| J | 896 hCoV-19/England/ALDP-94C7A0/2020\|EPI_ISL_556053\|20 | ···················· | ···················· | ····················· | ···················· | ···················· | ···················· |
| J | 897 hCoV-19/England/ALDP-94C5F1/2020\|EPI_ISL_556054\|20 | ···················· | ···················· | ····················· | ···················· | ···················· | ···················· |
| J | 898 hCoV-19/England/ALDP-94CCED/2020\|EPI_ISL_556055\|20 | ···················· | ·A·················· | ····················· | ···················· | ···················· | ···················· |
| J | 899 hCoV-19/England/ALDP-94CE8D/2020\|EPI_ISL_556056\|20 | ···················· | ···················· | ····················· | ···················· | ···················· | ···················· |
| J | 900 hCoV-19/England/ALDP-94C791/2020\|EPI_ISL_556057\|20 | ···················· | ···················· | ····················· | ···················· | ···················· | ···················· |
| J | 901 hCoV-19/England/CAMC-948F06/2020\|EPI_ISL_556058\|20 | ···················· | ···················· | ····················· | ···················· | ···················· | ···················· |
| J | 902 hCoV-19/England/ALDP-94C5E2/2020\|EPI_ISL_556059\|20 | ···················· | ···················· | ····················· | ···················· | ···················· | ···················· |
| J | 903 hCoV-19/England/ALDP-94F37C/2020\|EPI_ISL_556060\|20 | ···················· | ···················· | ····················· | ···················· | ···················· | ···················· |
| J | 904 hCoV-19/England/ALDP-94C755/2020\|EPI_ISL_556061\|20 | ···················· | ···················· | ····················· | ···················· | ···················· | ···················· |
| J | 905 hCoV-19/England/ALDP-94C97D/2020\|EPI_ISL_556062\|20 | ···················· | ···················· | ····················· | ···················· | ···················· | ···················· |
| J | 906 hCoV-19/England/CAMC-9496A6/2020\|EPI_ISL_556063\|20 | ···················· | ···················· | ····················· | ···················· | ···················· | ···················· |
| J | 907 hCoV-19/England/ALDP-94F3E5/2020\|EPI_ISL_556064\|20 | ···················· | ···················· | ····················· | ···················· | ···················· | ···················· |
| J | 908 hCoV-19/England/ALDP-94D420/2020\|EPI_ISL_556065\|20 | ···················· | ···················· | ···········T········· | ···················· | ···················· | ···················· |
| J | 909 hCoV-19/England/ALDP-94C870/2020\|EPI_ISL_556067\|20 | ···················· | ···················· | ····················· | ···················· | ···················· | ···················· |
| J | 910 hCoV-19/England/ALDP-94CBFF/2020\|EPI_ISL_556068\|20 | ···················· | ···················· | ····················· | ···················· | ···················· | ···················· |
| J | 911 hCoV-19/England/MILK-8184B6/2020\|EPI_ISL_557300\|20 | ···················· | ···················· | ····················· | ···················· | ···················· | ···················· |
| J | 912 hCoV-19/England/MILK-83982C/2020\|EPI_ISL_557303\|20 | ···················· | ···················· | ····················· | ···················· | ···················· | ···················· |
| J | 913 hCoV-19/England/MILK-839CE4/2020\|EPI_ISL_557305\|20 | ···················· | ···················· | ····················· | ···················· | ···················· | ···················· |
| J | 914 hCoV-19/England/MILK-80900F/2020\|EPI_ISL_557307\|20 | ···················· | ···················· | ····················· | ···················· | ···················· | ···················· |
| J | 915 hCoV-19/England/MILK-84A73A/2020\|EPI_ISL_557309\|20 | ···················· | ···················· | ····················· | ···················· | ···················· | ···················· |
| J | 916 hCoV-19/England/MILK-843D20/2020\|EPI_ISL_557310\|20 | ···················· | ···················· | ····················· | ···················· | ···················· | ···················· |
| J | 917 hCoV-19/England/MILK-84F8BA/2020\|EPI_ISL_557311\|20 | ···················· | ···················· | ····················· | ···················· | ···················· | ···················· |
| J | 918 hCoV-19/England/MILK-855C79/2020\|EPI_ISL_557312\|20 | ···················· | ·····A·············· | ····················· | ···················· | ···················· | ···················· |
| J | 919 hCoV-19/England/MILK-822D18/2020\|EPI_ISL_557314\|20 | ···················· | ···················· | ····················· | ···················· | ···················· | ···················· |
| J | 920 hCoV-19/England/MILK-82FE72/2020\|EPI_ISL_557315\|20 | ···················· | ···················· | ····················· | ···················· | ···················· | ···················· |
| J | 921 hCoV-19/England/MILK-809C1A/2020\|EPI_ISL_557316\|20 | ···················· | ·A·················· | ····················· | ···················· | ···················· | ···················· |
| J | 922 hCoV-19/England/ALDP-6FC72A/2020\|EPI_ISL_557380\|20 | ···················· | ···················· | ····················· | ···················· | ···················· | ···················· |
| J | 923 hCoV-19/England/ALDP-6FBB01/2020\|EPI_ISL_557381\|20 | ···················· | ···················· | ····················· | ···················· | ···················· | ···················· |
| J | 924 hCoV-19/England/MILK-85E3E4/2020\|EPI_ISL_557384\|20 | ···················· | ···················· | ····················· | ···················· | ···················· | ···················· |
| J | 925 hCoV-19/England/MILK-74A6A7/2020\|EPI_ISL_557385\|20 | ···················· | ···················· | ····················· | ···················· | ···················· | ···················· |
| J | 926 hCoV-19/England/MILK-6F81D5/2020\|EPI_ISL_557386\|20 | ···················· | ···················· | ····················· | ···················· | ···················· | ···················· |
| J | 927 hCoV-19/England/MILK-851FFC/2020\|EPI_ISL_557387\|20 | ···················· | ···················· | ····················· | ···················· | ···················· | ···················· |
| J | 928 hCoV-19/Wales/ALDP-6FA874/2020\|EPI_ISL_557388\|2020 | ···················· | ···················· | ····················· | ···················· | ···················· | ···················· |
| J | 929 hCoV-19/England/MILK-74EE41/2020\|EPI_ISL_557389\|20 | ···················· | ···················· | ····················· | ···················· | ···················· | ···················· |
| J | 930 hCoV-19/England/ALDP-6FAD0C/2020\|EPI_ISL_557390\|20 | ···················· | ···················· | ····················· | ···················· | ···················· | ···················· |
| J | 931 hCoV-19/England/MILK-8579E9/2020\|EPI_ISL_557391\|20 | ···················· | ···················· | ····················· | ···················· | ···················· | ···················· |
| J | 932 hCoV-19/England/MILK-8664F5/2020\|EPI_ISL_557393\|20 | ···················· | ···················· | ····················· | ···················· | ···················· | ···················· |
| J | 933 hCoV-19/England/MILK-874069/2020\|EPI_ISL_557394\|20 | ···················· | ···················· | ····················· | ···················· | ···················· | ···················· |
| J | 934 hCoV-19/England/MILK-7521AB/2020\|EPI_ISL_557395\|20 | ···················· | ···················· | ····················· | ···················· | ···················· | ···················· |
| J | 935 hCoV-19/England/MILK-8522E3/2020\|EPI_ISL_557396\|20 | ···················· | ·····A·············· | ····················· | ···················· | ···················· | ···················· |

```
J     936 hCoV-19/England/MILK-8523A4/2020|EPI_ISL_557397|20   ····················  ······A·············  ·····················  ····················  ····················  ····················
J     937 hCoV-19/England/ALDP-745A82/2020|EPI_ISL_557398|20   ····················  ····················  ·····················  ····················  ····················  ····················
J     938 hCoV-19/England/ALDP-6FCA6D/2020|EPI_ISL_557399|20   ····················  ····················  ·····················  ····················  ····················  ····················
J     939 hCoV-19/England/ALDP-5CAC18/2020|EPI_ISL_558600|20   ····················  ····················  ·····················  ····················  ····················  ····················
J     940 hCoV-19/England/ALDP-5CA313/2020|EPI_ISL_558602|20   ····················  ····················  ·····················  ····················  ····················  ····················
J     941 hCoV-19/England/ALDP-5C746E/2020|EPI_ISL_558603|20   ····················  ····················  ·····················  ····················  ····················  ····················
J     942 hCoV-19/England/ALDP-5CA97B/2020|EPI_ISL_558604|20   ····················  ····················  ·····················  ····················  ····················  ····················
J     943 hCoV-19/England/ALDP-5CB8B9/2020|EPI_ISL_558606|20   ····················  ····················  ·····················  ····················  ····················  ····················
J     944 hCoV-19/England/ALDP-5C9F5C/2020|EPI_ISL_558607|20   ····················  ····················  ·····················  ····················  ····················  ····················
J     945 hCoV-19/England/ALDP-5D9706/2020|EPI_ISL_558608|20   ····················  ····················  ·····················  ····················  ····················  ····················
J     946 hCoV-19/England/ALDP-5CD34D/2020|EPI_ISL_558610|20   ····················  ····················  ·····················  ····················  ····················  ····················
J     947 hCoV-19/England/ALDP-5D2D38/2020|EPI_ISL_558611|20   ····················  ····················  ·····················  ····················  ····················  ····················
J     948 hCoV-19/England/ALDP-5CAFA6/2020|EPI_ISL_558612|20   ····················  ····················  ·····················  ····················  ····················  ····················
J     949 hCoV-19/England/ALDP-5CFC04/2020|EPI_ISL_558613|20   ····················  ····················  ·····················  ····················  ····················  ····················
J     950 hCoV-19/England/ALDP-5CF466/2020|EPI_ISL_558614|20   ····················  ····················  ·····················  ····················  ····················  ····················
J     951 hCoV-19/England/ALDP-5CF9EF/2020|EPI_ISL_558615|20   ····················  ····················  ·····················  ····················  ····················  ····················
J     952 hCoV-19/England/ALDP-5C94A8/2020|EPI_ISL_558616|20   ····················  ····················  ·····················  ····················  ····················  ····················
J     953 hCoV-19/England/ALDP-5C3ECE/2020|EPI_ISL_558618|20   ····················  ····················  ·····················  ····················  ····················  ····················
J     954 hCoV-19/England/ALDP-59E6B5/2020|EPI_ISL_558619|20   ····················  ····················  ·····················  ····················  ····················  ····················
J     955 hCoV-19/England/ALDP-5A3A0C/2020|EPI_ISL_558620|20   ····················  ····················  ·····················  ····················  ····················  ····················
J     956 hCoV-19/England/ALDP-5C12D3/2020|EPI_ISL_558621|20   ····················  ····················  ·····················  ····················  ····················  ····················
J     957 hCoV-19/England/ALDP-5B6D1C/2020|EPI_ISL_558622|20   ····················  ····················  ·····················  ····················  ····················  ····················
J     958 hCoV-19/England/ALDP-5A9ACA/2020|EPI_ISL_558623|20   ····················  ····················  ·····················  ····················  ····················  ····················
J     959 hCoV-19/Wales/ALDP-5C2102/2020|EPI_ISL_558625|2020   ····················  ····················  ·····················  ····················  ····················  ····················
J     960 hCoV-19/England/ALDP-5AE245/2020|EPI_ISL_558626|20   ····················  ····················  ·····················  ····················  ····················  ····················
J     961 hCoV-19/England/ALDP-5AA9EA/2020|EPI_ISL_558628|20   ····················  ····················  ·····················  ····················  ····················  ····················
J     962 hCoV-19/England/ALDP-5A55BE/2020|EPI_ISL_558629|20   ····················  ····················  ·····················  ····················  ····················  ····················
J     963 hCoV-19/England/ALDP-5A88EF/2020|EPI_ISL_558631|20   ····················  ····················  ·····················  ····················  ····················  ····················
J     964 hCoV-19/England/ALDP-5A9B7C/2020|EPI_ISL_558632|20   ····················  ····················  ·····················  ····················  ····················  ····················
J     965 hCoV-19/England/ALDP-5BE4E2/2020|EPI_ISL_558633|20   ····················  ·A··················  ·····················  ····················  ····················  ····················
J     966 hCoV-19/England/ALDP-5C16C8/2020|EPI_ISL_558634|20   ····················  ····················  ·····················  ····················  ····················  ····················
J     967 hCoV-19/England/ALDP-5A2CAD/2020|EPI_ISL_558635|20   ····················  ····················  ·····················  ····················  ····················  ····················
J     968 hCoV-19/England/ALDP-5A0656/2020|EPI_ISL_558636|20   ····················  ····················  ·····················  ····················  ····················  ····················
J     969 hCoV-19/England/ALDP-5A3F58/2020|EPI_ISL_558637|20   ····················  ····················  ·····················  ····················  ····················  ····················
J     970 hCoV-19/England/MILK-5715F7/2020|EPI_ISL_558638|20   ····················  ····················  ·····················  ····················  ····················  ····················
J     971 hCoV-19/England/ALDP-5A525D/2020|EPI_ISL_558639|20   ····················  ····················  ·····················  ····················  ····················  ····················
J     972 hCoV-19/Wales/ALDP-5A05A4/2020|EPI_ISL_558641|2020   ····················  ····················  ·····················  ····················  ····················  ····················
J     973 hCoV-19/England/ALDP-5B69FA/2020|EPI_ISL_558643|20   ····················  ····················  ·····················  ····················  ····················  ····················
J     974 hCoV-19/England/ALDP-5B0A57/2020|EPI_ISL_558644|20   ····················  ····················  ·····················  ····················  ····················  ····················
J     975 hCoV-19/England/ALDP-59CAD9/2020|EPI_ISL_558692|20   ····················  ····················  ·····················  ····················  ····················  ····················
J     976 hCoV-19/England/ALDP-5AA99F/2020|EPI_ISL_558693|20   ····················  ····················  ·····················  ····················  ····················  ····················
J     977 hCoV-19/England/ALDP-5A5F74/2020|EPI_ISL_558694|20   ····················  ····················  ·····················  ····················  ····················  ····················
J     978 hCoV-19/England/ALDP-5B1E2D/2020|EPI_ISL_558696|20   ····················  ·A··················  ·····················  ····················  ····················  ····················
J     979 hCoV-19/England/ALDP-5A2D22/2020|EPI_ISL_558697|20   ····················  ····················  ·····················  ····················  ····················  ····················
J     980 hCoV-19/England/ALDP-5A8E38/2020|EPI_ISL_558698|20   ····················  ····················  ·····················  ····················  ····················  ····················
J     981 hCoV-19/England/ALDP-59F06B/2020|EPI_ISL_558699|20   ····················  ····················  ·····················  ····················  ····················  ····················
J     982 hCoV-19/England/SHEF-C290F/2020|EPI_ISL_559901|202   ····················  ·A··················  ·····················  ····················  ····················  ···A················
J     983 hCoV-19/England/SHEF-C48F1/2020|EPI_ISL_559907|202   ····················  ····················  ·····················  ····················  ····················  ····················
J     984 hCoV-19/England/SHEF-C5F73/2020|EPI_ISL_559909|202   ····················  ····················  ·····················  ····················  ····················  ····················
J     985 hCoV-19/England/SHEF-C683B/2020|EPI_ISL_559910|202   ····················  ····················  ·····················  ····················  ····················  ····················
J     986 hCoV-19/England/SHEF-C69B0/2020|EPI_ISL_559912|202   ····················  ····················  ·····················  ····················  ····················  ····················
J     987 hCoV-19/England/SHEF-C6AF9/2020|EPI_ISL_559913|202   ····················  ····················  ·····················  ····················  ····················  ····················
J     988 hCoV-19/England/SHEF-C714E/2020|EPI_ISL_559916|202   ····················  ····················  ·····················  ····················  ····················  ····················
J     989 hCoV-19/England/SHEF-C7849/2020|EPI_ISL_559918|202   ····················  ····················  ·····················  ····················  ····················  ····················
J     990 hCoV-19/England/SHEF-C7DB3/2020|EPI_ISL_559920|202   ····················  ····················  ·····················  ····················  ····················  ····················
J     991 hCoV-19/England/SHEF-C8848/2020|EPI_ISL_559922|202   ····················  ····················  ·····················  ····················  ····················  ····················
J     992 hCoV-19/England/SHEF-C9C96/2020|EPI_ISL_559927|202   ····················  ····················  ·····················  ····················  ····················  ····················
J     993 hCoV-19/England/SHEF-CC1B2/2020|EPI_ISL_559930|202   ····················  ····················  ·····················  ····················  ····················  ····················
J     994 hCoV-19/England/SHEF-CC1EF/2020|EPI_ISL_559931|202   ····················  ····················  ·····················  ····················  ····················  ····················
J     995 hCoV-19/England/SHEF-CC5B6/2020|EPI_ISL_559932|202   ····················  ····················  ·····················  ····················  ····················  ····················
J     996 hCoV-19/England/SHEF-CD746/2020|EPI_ISL_559933|202   ····················  ····················  ·····················  ····················  ····················  ····················
J     997 hCoV-19/England/SHEF-CE350/2020|EPI_ISL_559934|202   ····················  ····················  ·····················  ····················  ····················  ····················
J     998 hCoV-19/England/SHEF-CE44E/2020|EPI_ISL_559935|202   ····················  ····················  ·····················  ····················  ····················  ····················
J     999 hCoV-19/England/SHEF-CE4F3/2020|EPI_ISL_559936|202   ····················  ·A··················  ·····················  ····················  ····················  ····················
J    1000 hCoV-19/England/SHEF-D1A64/2020|EPI_ISL_559941|202   ····················  ····················  ·····················  ····················  ····················  ····················
J    1001 hCoV-19/England/SHEF-D1EFF/2020|EPI_ISL_559943|202   ····················  ····················  ·····················  ····················  ····················  ····················
J    1002 hCoV-19/England/SHEF-D2B33/2020|EPI_ISL_559946|202   ····················  ····················  ·····················  ····················  ····················  ····················
J    1003 hCoV-19/Scotland/CVR4273/2020|EPI_ISL_559947|2020-   ····················  ····················  ·····················  ····················  ····················  ····················
J    1004 hCoV-19/Scotland/CVR4275/2020|EPI_ISL_559948|2020-   ····················  ····················  ·····················  ····················  ····················  ····················
J    1005 hCoV-19/Scotland/CVR4276/2020|EPI_ISL_559949|2020-   ····················  ····················  ·····················  ····················  ····················  ····················
J    1006 hCoV-19/Scotland/CVR4277/2020|EPI_ISL_559950|2020-   ····················  ····················  ·····················  ····················  ····················  ····················
J    1007 hCoV-19/Scotland/CVR4279/2020|EPI_ISL_559952|2020-   ····················  ····················  ·····················  ····················  ····················  ····················
J    1008 hCoV-19/Scotland/CVR4281/2020|EPI_ISL_559954|2020-   ····················  ····················  ·····················  ····················  ····················  ····················
J    1009 hCoV-19/Scotland/CVR4282/2020|EPI_ISL_559955|2020-   ····················  ····················  ·····················  ····················  ····················  ····················
J    1010 hCoV-19/Scotland/CVR4283/2020|EPI_ISL_559956|2020-   ····················  ····················  ·····················  ····················  ····················  ····················
J    1011 hCoV-19/Scotland/CVR4285/2020|EPI_ISL_559958|2020-   ····················  ····················  ·····················  ····················  ····················  ····················
J    1012 hCoV-19/Scotland/CVR4287/2020|EPI_ISL_559960|2020-   ····················  ····················  ·····················  ····················  ····················  ····················
J    1013 hCoV-19/Scotland/CVR4290/2020|EPI_ISL_559962|2020-   ····················  ····················  ·····················  ····················  ····················  ····················
J    1014 hCoV-19/Scotland/CVR4292/2020|EPI_ISL_559964|2020-   ····················  ····················  ·····················  ····················  ····················  ····················
J    1015 hCoV-19/Scotland/CVR4294/2020|EPI_ISL_559966|2020-   ····················  ····················  ·····················  ····················  ····················  ····················
```

| | | | | | | | |
|---|---|---|---|---|---|---|---|
| J | 1016 hCoV-19/Scotland/CVR4295/2020\|EPI_ISL_559967\|2020- | ···················· | ···················· | ···················· | ···················· | ···················· | ···················· |
| J | 1017 hCoV-19/Scotland/CVR4297/2020\|EPI_ISL_559968\|2020- | ···················· | ···················· | ···················· | ···················· | ···················· | ···················· |
| J | 1018 hCoV-19/Scotland/CVR4298/2020\|EPI_ISL_559969\|2020- | ···················· | ···················· | ···················· | ···················· | ···················· | ···················· |
| J | 1019 hCoV-19/Scotland/CVR4299/2020\|EPI_ISL_559970\|2020- | ···················· | ···················· | ···················· | ···················· | ···················· | ···················· |
| J | 1020 hCoV-19/Scotland/EDB1030/2020\|EPI_ISL_559971\|2020- | ···················· | ···················· | ···················· | ···················· | ···················· | ···················· |
| J | 1021 hCoV-19/Scotland/EDB1155/2020\|EPI_ISL_559972\|2020- | ···················· | ···················· | ···················· | ···················· | ···················· | ···················· |
| J | 1022 hCoV-19/Scotland/EDB3708/2020\|EPI_ISL_559974\|2020- | ···················· | ···················· | ···················· | ···················· | ···················· | ···················· |
| J | 1023 hCoV-19/Scotland/EDB6890/2020\|EPI_ISL_559976\|2020- | ···················· | ·A·················· | ···················· | ···················· | ···················· | ···················· |
| J | 1024 hCoV-19/Scotland/EDB6892/2020\|EPI_ISL_559978\|2020- | ···················· | ·A·················· | ···················· | ···················· | ···················· | ···················· |
| J | 1025 hCoV-19/England/MILK-9A957A/2020\|EPI_ISL_551202\|20 | ···················· | ···················· | ···················· | ···················· | ···················· | ···················· |
| J | 1026 hCoV-19/England/MILK-9A99C9/2020\|EPI_ISL_551205\|20 | ···················· | ···················· | ···················· | ···················· | ···················· | ···················· |
| J | 1027 hCoV-19/England/MILK-9AAE6F/2020\|EPI_ISL_551206\|20 | ···················· | ···················· | ···················· | ···················· | ······T············· | ···················· |
| J | 1028 hCoV-19/England/MILK-9A9334/2020\|EPI_ISL_551207\|20 | ···················· | ···················· | ···················· | ···················· | ···················· | ···············A···· |
| J | 1029 hCoV-19/England/MILK-9AAE23/2020\|EPI_ISL_551208\|20 | ···················· | ···················· | ···················· | ···················· | ···················· | ···················· |
| J | 1030 hCoV-19/England/MILK-9A9EBB/2020\|EPI_ISL_551209\|20 | ···················· | ···················· | ···················· | ···················· | ···················· | ···················· |
| J | 1031 hCoV-19/England/MILK-9AB095/2020\|EPI_ISL_551210\|20 | ···················· | ···················· | ···················· | ···················· | ···················· | ···················· |
| J | 1032 hCoV-19/England/MILK-9A8ECB/2020\|EPI_ISL_551258\|20 | ···················· | ···················· | ···················· | ···················· | ···················· | ···················· |
| J | 1033 hCoV-19/England/MILK-9AB3BA/2020\|EPI_ISL_551259\|20 | ···················· | ···················· | ···················· | ···················· | ···················· | ···················· |
| J | 1034 hCoV-19/England/MILK-9AAE8D/2020\|EPI_ISL_551267\|20 | ···················· | ···················· | ···················· | ···················· | ······T············· | ···················· |
| J | 1035 hCoV-19/England/MILK-9A9ECA/2020\|EPI_ISL_551268\|20 | ···················· | ···················· | ···················· | ···················· | ···················· | ···················· |
| J | 1036 hCoV-19/England/MILK-9A901F/2020\|EPI_ISL_551269\|20 | ···················· | ·A·················· | ···················· | ···················· | ···················· | ···················· |
| J | 1037 hCoV-19/England/MILK-9A9352/2020\|EPI_ISL_551270\|20 | ···················· | ·A·················· | ···················· | ···················· | ···················· | ···················· |
| J | 1038 hCoV-19/England/MILK-9AAEAB/2020\|EPI_ISL_551271\|20 | ···················· | ···················· | ···················· | ···················· | ···················· | ···················· |
| J | 1039 hCoV-19/England/MILK-9AAEC9/2020\|EPI_ISL_551272\|20 | ···················· | ·A·················· | ···················· | ···················· | ···················· | ···················· |
| J | 1040 hCoV-19/England/MILK-9A9CA2/2020\|EPI_ISL_551273\|20 | ···················· | ···················· | ···················· | ···················· | ···················· | ···················· |
| J | 1041 hCoV-19/England/MILK-9A9E24/2020\|EPI_ISL_551274\|20 | ···················· | ···················· | ···················· | ···················· | ···················· | ···················· |
| J | 1042 hCoV-19/England/MILK-9A9237/2020\|EPI_ISL_551277\|20 | ···················· | ···················· | ···················· | ···················· | ···················· | ···················· |
| J | 1043 hCoV-19/England/MILK-9A8E52/2020\|EPI_ISL_551278\|20 | ···················· | ···················· | ···················· | ···················· | ···················· | ···················· |
| J | 1044 hCoV-19/England/MILK-9A938F/2020\|EPI_ISL_551279\|20 | ···················· | ···················· | ···················· | ···················· | ···················· | ···················· |
| J | 1045 hCoV-19/England/MILK-9A9D09/2020\|EPI_ISL_551280\|20 | ···················· | ···················· | ···················· | ···················· | ···················· | ···················· |
| J | 1046 hCoV-19/England/MILK-9A9DDC/2020\|EPI_ISL_551281\|20 | ···················· | ·A·················· | ···················· | ···················· | ···················· | ···················· |
| J | 1047 hCoV-19/England/MILK-9A92A0/2020\|EPI_ISL_551284\|20 | ···················· | ···················· | ···················· | ···················· | ···················· | ···················· |
| J | 1048 hCoV-19/England/ALDP-951BBB/2020\|EPI_ISL_551285\|20 | ···················· | ···················· | ···················· | ···················· | ···················· | ···················· |
| J | 1049 hCoV-19/England/ALDP-951F55/2020\|EPI_ISL_551289\|20 | ···················· | ···················· | ···················· | ···················· | ···················· | ···················· |
| J | 1050 hCoV-19/England/ALDP-952288/2020\|EPI_ISL_551290\|20 | ···················· | ···················· | ···················· | ···················· | ···················· | ···················· |
| J | 1051 hCoV-19/England/ALDP-951DE2/2020\|EPI_ISL_551291\|20 | ···················· | ···················· | ···················· | ···················· | ···················· | ···················· |
| J | 1052 hCoV-19/England/ALDP-9522B5/2020\|EPI_ISL_551292\|20 | ···················· | ···················· | ···················· | ···················· | ···················· | ···················· |
| J | 1053 hCoV-19/England/ALDP-951E0D/2020\|EPI_ISL_551293\|20 | ···················· | ···················· | ···················· | ···················· | ···················· | ···················· |
| J | 1054 hCoV-19/England/ALDP-952006/2020\|EPI_ISL_551295\|20 | ···················· | ···················· | ···················· | ···················· | ···················· | ···················· |
| J | 1055 hCoV-19/England/ALDP-9522E2/2020\|EPI_ISL_551296\|20 | ···················· | ···················· | ···················· | ···················· | ···················· | ···················· |
| J | 1056 hCoV-19/England/ALDP-951C4F/2020\|EPI_ISL_551298\|20 | ···················· | ···················· | ···················· | ···················· | ···················· | ···················· |
| J | 1057 hCoV-19/England/ALDP-951B60/2020\|EPI_ISL_551299\|20 | ···················· | ···················· | ···················· | ···················· | ···················· | ···················· |
| J | 1058 hCoV-19/England/MILK-9754A7/2020\|EPI_ISL_552500\|20 | ···················· | ·A·················· | ···················· | ···················· | ···················· | ···A················ |
| J | 1059 hCoV-19/England/MILK-975B1B/2020\|EPI_ISL_552501\|20 | ···················· | ···················· | ···················· | ···················· | ···················· | ···················· |
| J | 1060 hCoV-19/England/MILK-97535F/2020\|EPI_ISL_552502\|20 | ···················· | ···················· | ···················· | ···················· | ···················· | ···················· |
| J | 1061 hCoV-19/England/MILK-974B67/2020\|EPI_ISL_552503\|20 | ···················· | ···················· | ···················· | ···················· | ···················· | ···················· |
| J | 1062 hCoV-19/England/MILK-9751CE/2020\|EPI_ISL_552504\|20 | ···················· | ···················· | ···················· | ···················· | ···················· | ···················· |
| J | 1063 hCoV-19/England/MILK-975A78/2020\|EPI_ISL_552505\|20 | ···················· | ···················· | ···················· | ···················· | ···················· | ···················· |
| J | 1064 hCoV-19/England/ALDP-982BA2/2020\|EPI_ISL_552506\|20 | ···················· | ···················· | ···················· | ···················· | ···················· | ···················· |
| J | 1065 hCoV-19/England/ALDP-9829F3/2020\|EPI_ISL_552507\|20 | ···················· | ···················· | ···················· | ···················· | ···················· | ···················· |
| J | 1066 hCoV-19/England/ALDP-982BFD/2020\|EPI_ISL_552508\|20 | ···················· | ···················· | ···················· | ···················· | ···················· | ···················· |
| J | 1067 hCoV-19/England/MILK-974C91/2020\|EPI_ISL_552509\|20 | ···················· | ···················· | ···················· | ···················· | ···················· | ···················· |
| J | 1068 hCoV-19/England/MILK-974D07/2020\|EPI_ISL_552510\|20 | ···················· | ···················· | ···················· | ···················· | ···················· | ···················· |
| J | 1069 hCoV-19/England/MILK-974D61/2020\|EPI_ISL_552511\|20 | ···················· | ···················· | ···················· | ···················· | ···················· | ···················· |
| J | 1070 hCoV-19/England/ALDP-982911/2020\|EPI_ISL_552512\|20 | ···················· | ···················· | ···················· | ···················· | ···················· | ···················· |
| J | 1071 hCoV-19/England/MILK-975128/2020\|EPI_ISL_552513\|20 | ···················· | ···················· | ···················· | ···················· | ···················· | ···················· |
| J | 1072 hCoV-19/England/ALDP-9828E7/2020\|EPI_ISL_552515\|20 | ···················· | ·A·················· | ···················· | ···················· | ···················· | ···················· |
| J | 1073 hCoV-19/England/ALDP-982A3C/2020\|EPI_ISL_552516\|20 | ···················· | ···················· | ···················· | ···················· | ···················· | ···················· |
| J | 1074 hCoV-19/England/MILK-974CBF/2020\|EPI_ISL_552517\|20 | ···················· | ···················· | ···················· | ···················· | ···················· | ···················· |
| J | 1075 hCoV-19/England/MILK-975137/2020\|EPI_ISL_552518\|20 | ···················· | ···················· | ···················· | ···················· | ···················· | ···················· |
| J | 1076 hCoV-19/England/MILK-974DDA/2020\|EPI_ISL_552520\|20 | ···················· | ···················· | ···················· | ···················· | ···················· | ···················· |
| J | 1077 hCoV-19/England/MILK-975568/2020\|EPI_ISL_552522\|20 | ···················· | ···················· | ···················· | ···················· | ···C················ | ···················· |
| J | 1078 hCoV-19/England/ALDP-982841/2020\|EPI_ISL_552523\|20 | ···················· | ···················· | ···················· | ···················· | ···················· | ···················· |
| J | 1079 hCoV-19/England/MILK-975762/2020\|EPI_ISL_552524\|20 | ···················· | ···················· | ···················· | ···················· | ···················· | ···················· |
| J | 1080 hCoV-19/England/MILK-974AF1/2020\|EPI_ISL_552525\|20 | ···················· | ···················· | ···················· | ···················· | ···················· | ···················· |
| J | 1081 hCoV-19/England/ALDP-982A4B/2020\|EPI_ISL_552526\|20 | ···················· | ···················· | ···················· | ···················· | ···················· | ···················· |
| J | 1082 hCoV-19/England/ALDP-9828C9/2020\|EPI_ISL_552527\|20 | ···················· | ···················· | ···················· | ···················· | ···················· | ···················· |
| J | 1083 hCoV-19/England/MILK-974CDD/2020\|EPI_ISL_552528\|20 | ···················· | ···················· | ···················· | ···················· | ···················· | ···················· |
| J | 1084 hCoV-19/England/MILK-975C45/2020\|EPI_ISL_552529\|20 | ···················· | ···················· | ···················· | ···················· | ···················· | ···················· |
| J | 1085 hCoV-19/England/ALDP-982A96/2020\|EPI_ISL_552530\|20 | ···················· | ···················· | ···················· | ···················· | ···················· | ···················· |
| J | 1086 hCoV-19/England/MILK-974E8C/2020\|EPI_ISL_552531\|20 | ···················· | ···················· | ···················· | ···················· | ···················· | ···················· |
| J | 1087 hCoV-19/England/ALDP-982A1E/2020\|EPI_ISL_552533\|20 | ···················· | ···················· | ···················· | ···················· | ···················· | ···················· |
| J | 1088 hCoV-19/England/MILK-974D8F/2020\|EPI_ISL_552535\|20 | ···················· | ···················· | ···················· | ···················· | ···················· | ···················· |
| J | 1089 hCoV-19/England/MILK-975717/2020\|EPI_ISL_552536\|20 | ···················· | ···················· | ···················· | ···················· | ···················· | ···················· |
| J | 1090 hCoV-19/England/MILK-975401/2020\|EPI_ISL_552538\|20 | ···················· | ·········A·········· | ···················· | ···················· | ······T············· | ···················· |
| J | 1091 hCoV-19/England/ALDP-982B39/2020\|EPI_ISL_552539\|20 | ···················· | ·A·················· | ···················· | ···················· | ···················· | ···A················ |
| J | 1092 hCoV-19/England/ALDP-982B93/2020\|EPI_ISL_552540\|20 | ···················· | ···················· | ···················· | ···················· | ···················· | ···················· |
| J | 1093 hCoV-19/England/MILK-9758D8/2020\|EPI_ISL_552541\|20 | ···················· | ···················· | ···················· | ···················· | ···················· | ···················· |
| J | 1094 hCoV-19/England/MILK-974C73/2020\|EPI_ISL_552542\|20 | ···················· | ···················· | ···················· | ···················· | ···················· | ···················· |
| J | 1095 hCoV-19/England/ALDP-982AE1/2020\|EPI_ISL_552587\|20 | ···················· | ···················· | ···················· | ···················· | ···················· | ···················· |

| | | | | | | | | |
|---|---|---|---|---|---|---|---|---|
| J | 1096 | hCoV-19/England/MILK-974B0D/2020\|EPI_ISL_552588\|20 | ···················· | ···················· | ···················· | ···················· | ···················· | ···················· |
| J | 1097 | hCoV-19/England/MILK-974BA3/2020\|EPI_ISL_552589\|20 | ···················· | ···················· | ···················· | ···················· | ···················· | ···················· |
| J | 1098 | hCoV-19/England/ALDP-982CBE/2020\|EPI_ISL_552598\|20 | ···················· | ·A·················· | ···················· | ···················· | ···················· | ···A················ |
| J | 1099 | hCoV-19/England/MILK-9751A0/2020\|EPI_ISL_552599\|20 | ···················· | ···A················ | ···················· | ···················· | ···················· | ···················· |
| J | 1100 | hCoV-19/England/ALDP-9538B2/2020\|EPI_ISL_553800\|20 | ···················· | ···················· | ···················· | ···················· | ···················· | ···················· |
| J | 1101 | hCoV-19/England/ALDP-953937/2020\|EPI_ISL_553801\|20 | ···················· | ···················· | ···················· | ···················· | ···················· | ···················· |
| J | 1102 | hCoV-19/England/ALDP-954A60/2020\|EPI_ISL_553802\|20 | ···················· | ···················· | ···················· | ···················· | ···················· | ···················· |
| J | 1103 | hCoV-19/England/ALDP-95424A/2020\|EPI_ISL_553804\|20 | ···················· | ···················· | ···················· | ···················· | ···················· | ···················· |
| J | 1104 | hCoV-19/England/ALDP-953928/2020\|EPI_ISL_553805\|20 | ···················· | ···················· | ···················· | ···················· | ···················· | ···················· |
| J | 1105 | hCoV-19/England/ALDP-953F44/2020\|EPI_ISL_553806\|20 | ···················· | ···················· | ···················· | ···················· | ···················· | ···················· |
| J | 1106 | hCoV-19/England/ALDP-9540E6/2020\|EPI_ISL_553807\|20 | ···················· | ···················· | ···················· | ···················· | ···················· | ···················· |
| J | 1107 | hCoV-19/England/ALDP-953C10/2020\|EPI_ISL_553808\|20 | ···················· | ···················· | ···················· | ···················· | ···················· | ···················· |
| J | 1108 | hCoV-19/England/ALDP-953A70/2020\|EPI_ISL_553809\|20 | ···················· | ···················· | ···················· | ···················· | ···················· | ···················· |
| J | 1109 | hCoV-19/England/ALDP-95406E/2020\|EPI_ISL_553813\|20 | ···················· | ···················· | ···················· | ···················· | ···················· | ···················· |
| J | 1110 | hCoV-19/England/ALDP-953ECF/2020\|EPI_ISL_553814\|20 | ···················· | ···················· | ···················· | ···················· | ···················· | ···················· |
| J | 1111 | hCoV-19/England/ALDP-9534EB/2020\|EPI_ISL_553815\|20 | ···················· | ···················· | ···················· | ···················· | ···················· | ···················· |
| J | 1112 | hCoV-19/England/ALDP-95382B/2020\|EPI_ISL_553816\|20 | ···················· | ···················· | ···················· | ···················· | ···················· | ···················· |
| J | 1113 | hCoV-19/Wales/ALDP-954383/2020\|EPI_ISL_553819\|2020 | ···················· | ···················· | ···················· | ···················· | ···················· | ···················· |
| J | 1114 | hCoV-19/England/ALDP-9535E8/2020\|EPI_ISL_553820\|20 | ···················· | ···················· | ···················· | ···················· | ···················· | ···················· |
| J | 1115 | hCoV-19/England/ALDP-9542E0/2020\|EPI_ISL_553821\|20 | ···················· | ···················· | ···················· | ···················· | ···················· | ···················· |
| J | 1116 | hCoV-19/England/ALDP-9544DB/2020\|EPI_ISL_553822\|20 | ···················· | ···················· | ···················· | ···················· | ···················· | ···················· |
| J | 1117 | hCoV-19/England/ALDP-9541A7/2020\|EPI_ISL_553823\|20 | ···················· | ···················· | ···················· | ···················· | ···················· | ···················· |
| J | 1118 | hCoV-19/England/ALDP-953533/2020\|EPI_ISL_553824\|20 | ···················· | ···················· | ···················· | ···················· | ···················· | ···················· |
| J | 1119 | hCoV-19/England/ALDP-9538C1/2020\|EPI_ISL_553825\|20 | ···················· | ···················· | ···················· | ···················· | ···················· | ···················· |
| J | 1120 | hCoV-19/England/ALDP-95359D/2020\|EPI_ISL_553826\|20 | ···················· | ···················· | ···················· | ···················· | ···················· | ···················· |
| J | 1121 | hCoV-19/Wales/ALDP-9543B0/2020\|EPI_ISL_553827\|2020 | ···················· | ···················· | ···················· | ···················· | ···················· | ···················· |
| J | 1122 | hCoV-19/England/ALDP-9540F5/2020\|EPI_ISL_553828\|20 | ···················· | ···················· | ···················· | ···················· | ···················· | ···················· |
| J | 1123 | hCoV-19/England/ALDP-953885/2020\|EPI_ISL_553829\|20 | ···················· | ···················· | ···················· | ···················· | ···················· | ···················· |
| J | 1124 | hCoV-19/England/ALDP-954101/2020\|EPI_ISL_553830\|20 | ···················· | ···················· | ···················· | ···················· | ···················· | ···················· |
| J | 1125 | hCoV-19/England/ALDP-9538D0/2020\|EPI_ISL_553832\|20 | ···················· | ···················· | ···················· | ···················· | ···················· | ···················· |
| J | 1126 | hCoV-19/England/ALDP-95430B/2020\|EPI_ISL_553834\|20 | ···················· | ···················· | ···················· | ···················· | ···················· | ···················· |
| J | 1127 | hCoV-19/England/ALDP-953F17/2020\|EPI_ISL_553836\|20 | ···················· | ···················· | ···················· | ···················· | ···················· | ···················· |
| J | 1128 | hCoV-19/England/ALDP-95381C/2020\|EPI_ISL_553837\|20 | ···················· | ···················· | ···················· | ···················· | ···················· | ···················· |
| J | 1129 | hCoV-19/England/ALDP-95371F/2020\|EPI_ISL_553838\|20 | ···················· | ···················· | ···················· | ···················· | ···················· | ···················· |
| J | 1130 | hCoV-19/England/ALDP-9535CA/2020\|EPI_ISL_553840\|20 | ···················· | ···················· | ···················· | ···················· | ···················· | ···················· |
| J | 1131 | hCoV-19/England/ALDP-954365/2020\|EPI_ISL_553842\|20 | ···················· | ···················· | ···················· | ···················· | ···················· | ···················· |
| J | 1132 | hCoV-19/England/ALDP-953F35/2020\|EPI_ISL_553843\|20 | ···················· | ···················· | ···················· | ···················· | ···················· | ···················· |
| J | 1133 | hCoV-19/Wales/ALDP-9543A1/2020\|EPI_ISL_553844\|2020 | ···················· | ···················· | ···················· | ···················· | ···················· | ···················· |
| J | 1134 | hCoV-19/England/ALDP-95414D/2020\|EPI_ISL_553845\|20 | ···················· | ···················· | ···················· | ···················· | ···················· | ···················· |
| J | 1135 | hCoV-19/England/ALDP-9539DD/2020\|EPI_ISL_553847\|20 | ···················· | ···················· | ···················· | ···················· | ···················· | ···················· |
| J | 1136 | hCoV-19/England/ALDP-9535BB/2020\|EPI_ISL_553848\|20 | ···················· | ···················· | ···················· | ···················· | ···················· | ···················· |
| J | 1137 | hCoV-19/England/ALDP-9534FA/2020\|EPI_ISL_553849\|20 | ···················· | ···················· | ···················· | ···················· | ···················· | ···················· |
| J | 1138 | hCoV-19/England/ALDP-953C3E/2020\|EPI_ISL_553850\|20 | ···················· | ···················· | ···················· | ···················· | ···················· | ···················· |
| J | 1139 | hCoV-19/England/ALDP-953A16/2020\|EPI_ISL_553852\|20 | ···················· | ···················· | ···················· | ···················· | ···················· | ···················· |
| J | 1140 | hCoV-19/England/ALDP-95407D/2020\|EPI_ISL_553853\|20 | ···················· | ···················· | ···················· | ···················· | ···················· | ···················· |
| J | 1141 | hCoV-19/England/ALDP-954611/2020\|EPI_ISL_553855\|20 | ···················· | ···················· | ···················· | ···················· | ···················· | ···················· |
| J | 1142 | hCoV-19/England/ALDP-9537B5/2020\|EPI_ISL_553856\|20 | ···················· | ···················· | ···················· | ···················· | ···················· | ···················· |
| J | 1143 | hCoV-19/England/ALDP-95417A/2020\|EPI_ISL_553857\|20 | ···················· | ···················· | ···················· | ···················· | ···················· | ···················· |
| J | 1144 | hCoV-19/England/ALDP-95408C/2020\|EPI_ISL_553858\|20 | ···················· | ···················· | ···················· | ···················· | ···················· | ···················· |
| J | 1145 | hCoV-19/England/ALDP-954453/2020\|EPI_ISL_553859\|20 | ···················· | ···················· | ···················· | ···················· | ···················· | ···················· |
| J | 1146 | hCoV-19/England/ALDP-95372E/2020\|EPI_ISL_553860\|20 | ···················· | ···················· | ···················· | ···················· | ···················· | ···················· |
| J | 1147 | hCoV-19/England/ALDP-9539FB/2020\|EPI_ISL_553862\|20 | ···················· | ···················· | ···················· | ···················· | ···················· | ···················· |
| J | 1148 | hCoV-19/England/ALDP-9542FF/2020\|EPI_ISL_553863\|20 | ···················· | ···················· | ···················· | ···················· | ···················· | ···················· |
| J | 1149 | hCoV-19/England/ALDP-953C01/2020\|EPI_ISL_553864\|20 | ···················· | ···················· | ···················· | ···················· | ···················· | ···················· |
| J | 1150 | hCoV-19/England/ALDP-9535F7/2020\|EPI_ISL_553867\|20 | ···················· | ···················· | ···················· | ···················· | ···················· | ···················· |
| J | 1151 | hCoV-19/England/ALDP-953BF5/2020\|EPI_ISL_553869\|20 | ···················· | ···················· | ···················· | ···················· | ···················· | ···················· |
| J | 1152 | hCoV-19/England/ALDP-95463F/2020\|EPI_ISL_553870\|20 | ···················· | ···················· | ···················· | ···················· | ···················· | ···················· |
| J | 1153 | hCoV-19/England/ALDP-95610F/2020\|EPI_ISL_553872\|20 | ···················· | ···················· | ···················· | ···················· | ···················· | ···················· |
| J | 1154 | hCoV-19/England/ALDP-955BD5/2020\|EPI_ISL_553873\|20 | ···················· | ···················· | ···················· | ···················· | ···················· | ···················· |
| J | 1155 | hCoV-19/England/ALDP-94CD80/2020\|EPI_ISL_556070\|20 | ···················· | ···················· | ···················· | ···················· | ···················· | ···················· |
| J | 1156 | hCoV-19/England/ALDP-94C56A/2020\|EPI_ISL_556071\|20 | ···················· | ···················· | ···················· | ···················· | ···················· | ···················· |
| J | 1157 | hCoV-19/Wales/ALDP-94CA10/2020\|EPI_ISL_556072\|2020 | ···················· | ···················· | ···················· | ···················· | ···················· | ···················· |
| J | 1158 | hCoV-19/England/ALDP-94CFD5/2020\|EPI_ISL_556073\|20 | ···················· | ···················· | ···················· | ···················· | ···················· | ···················· |
| J | 1159 | hCoV-19/England/ALDP-6D9161/2020\|EPI_ISL_556074\|20 | ···················· | ···················· | ···················· | ···················· | ···················· | ···················· |
| J | 1160 | hCoV-19/England/MILK-95DA67/2020\|EPI_ISL_556075\|20 | ···················· | ···················· | ···················· | ···················· | ···················· | ···················· |
| J | 1161 | hCoV-19/England/MILK-95D997/2020\|EPI_ISL_556076\|20 | ···················· | ···················· | ···················· | ···················· | ···················· | ···················· |
| J | 1162 | hCoV-19/England/MILK-95D86D/2020\|EPI_ISL_556077\|20 | ···················· | ···················· | ···················· | ···················· | ···················· | ···················· |
| J | 1163 | hCoV-19/England/ALDP-6D5DE9/2020\|EPI_ISL_556078\|20 | ···················· | ···················· | ···················· | ···················· | ···················· | ···················· |
| J | 1164 | hCoV-19/England/MILK-95D85E/2020\|EPI_ISL_556079\|20 | ···················· | ···················· | ···················· | ···················· | ···················· | ···················· |
| J | 1165 | hCoV-19/England/ALDP-6C5414/2020\|EPI_ISL_556080\|20 | ···················· | ···················· | ···················· | ···················· | ···················· | ···················· |
| J | 1166 | hCoV-19/England/ALDP-6CC1AD/2020\|EPI_ISL_556081\|20 | ···················· | ···················· | ···················· | ···················· | ···················· | ···················· |
| J | 1167 | hCoV-19/England/ALDP-6CE475/2020\|EPI_ISL_556082\|20 | ···················· | ···················· | ···················· | ···················· | ···················· | ···················· |
| J | 1168 | hCoV-19/Wales/MILK-95DA58/2020\|EPI_ISL_556083\|2020 | ···················· | ···················· | ···················· | ···················· | ···················· | ···················· |
| J | 1169 | hCoV-19/England/ALDP-6CC7F6/2020\|EPI_ISL_556084\|20 | ···················· | ···················· | ···················· | ···················· | ···················· | ···················· |
| J | 1170 | hCoV-19/England/ALDP-94EEF4/2020\|EPI_ISL_556085\|20 | ···················· | ···················· | ···················· | ···················· | ···················· | ···················· |
| J | 1171 | hCoV-19/England/ALDP-6D08ED/2020\|EPI_ISL_556086\|20 | ···················· | ···················· | ···················· | ···················· | ···················· | ···················· |
| J | 1172 | hCoV-19/England/MILK-95DAFE/2020\|EPI_ISL_556087\|20 | ···················· | ···················· | ···················· | ···················· | ···················· | ···················· |
| J | 1173 | hCoV-19/Wales/ALDP-6CE967/2020\|EPI_ISL_556088\|2020 | ···················· | ···················· | ···················· | ···················· | ···················· | ···················· |
| J | 1174 | hCoV-19/England/ALDP-6CAC62/2020\|EPI_ISL_556089\|20 | ···················· | ···················· | ···················· | ···················· | ···················· | ···················· |
| J | 1175 | hCoV-19/England/MILK-95DAD0/2020\|EPI_ISL_556090\|20 | ···················· | ···················· | ···················· | ···················· | ···················· | ···················· |

| | | | | | | | | |
|---|---|---|---|---|---|---|---|---|
| J | 1176 | hCoV-19/England/MILK-95D89A/2020\|EPI_ISL_556092\|20 | ···················· | ···················· | ···················· | ···················· | ···················· | ···················· |
| J | 1177 | hCoV-19/England/MILK-631DBE/2020\|EPI_ISL_556093\|20 | ···················· | ···················· | ···················· | ···················· | ···················· | ···················· |
| J | 1178 | hCoV-19/England/MILK-95DB91/2020\|EPI_ISL_556095\|20 | ···················· | ···················· | ···················· | ···················· | ···················· | ···················· |
| J | 1179 | hCoV-19/England/MILK-95D7CA/2020\|EPI_ISL_556097\|20 | ···················· | ···················· | ···················· | ···················· | ···················· | ···················· |
| J | 1180 | hCoV-19/England/ALDP-6D56DF/2020\|EPI_ISL_556099\|20 | ···················· | ···················· | ···················· | ···················· | ···················· | ···················· |
| J | 1181 | hCoV-19/England/MILK-846C02/2020\|EPI_ISL_557202\|20 | ···················· | ···················· | ···················· | ···················· | ···················· | ···················· |
| J | 1182 | hCoV-19/England/MILK-83CD39/2020\|EPI_ISL_557203\|20 | ···················· | ···················· | ···················· | ···················· | ···················· | ···················· |
| J | 1183 | hCoV-19/England/MILK-83E64B/2020\|EPI_ISL_557204\|20 | ···················· | ···················· | ···················· | ···················· | ···················· | ···················· |
| J | 1184 | hCoV-19/England/MILK-80E811/2020\|EPI_ISL_557205\|20 | ···················· | ···················· | ···················· | ···················· | ···················· | ···················· |
| J | 1185 | hCoV-19/England/MILK-837C04/2020\|EPI_ISL_557206\|20 | ···················· | ···················· | ···················· | ···················· | ···················· | ···················· |
| J | 1186 | hCoV-19/England/MILK-8127E0/2020\|EPI_ISL_557208\|20 | ···················· | ···················· | ···················· | ···················· | ···················· | ···················· |
| J | 1187 | hCoV-19/England/MILK-80A29F/2020\|EPI_ISL_557209\|20 | ···················· | ···················· | ···················· | ···················· | ···················· | ···················· |
| J | 1188 | hCoV-19/England/MILK-82E3ED/2020\|EPI_ISL_557210\|20 | ···T················ | ···················· | ···················· | ···················· | ···················· | ···················· |
| J | 1189 | hCoV-19/England/MILK-8274D2/2020\|EPI_ISL_557211\|20 | ···T················ | ···················· | ···················· | ···················· | ···················· | ···················· |
| J | 1190 | hCoV-19/England/MILK-828A57/2020\|EPI_ISL_557212\|20 | ···················· | ·A·················· | ···················· | ···················· | ···················· | ···················· |
| J | 1191 | hCoV-19/England/MILK-81D527/2020\|EPI_ISL_557214\|20 | ···················· | ···················· | ···················· | ···················· | ···················· | ···················· |
| J | 1192 | hCoV-19/England/MILK-81E896/2020\|EPI_ISL_557215\|20 | ···················· | ······A············· | ···················· | ···················· | ···················· | ···················· |
| J | 1193 | hCoV-19/England/MILK-87A35B/2020\|EPI_ISL_557216\|20 | ···················· | ···················· | ···················· | ···················· | ···················· | ···················· |
| J | 1194 | hCoV-19/England/MILK-817D52/2020\|EPI_ISL_557217\|20 | ···················· | ···················· | ···················· | ···················· | ···················· | ···················· |
| J | 1195 | hCoV-19/England/MILK-84812F/2020\|EPI_ISL_557218\|20 | ···················· | ···················· | ···················· | ···················· | ···················· | ···················· |
| J | 1196 | hCoV-19/England/MILK-8174B7/2020\|EPI_ISL_557219\|20 | ···················· | ······A············· | ···················· | ···················· | ···················· | ···················· |
| J | 1197 | hCoV-19/England/MILK-82310C/2020\|EPI_ISL_557220\|20 | ···················· | ···················· | ···················· | ···················· | ···················· | ···················· |
| J | 1198 | hCoV-19/England/MILK-832823/2020\|EPI_ISL_557221\|20 | ···················· | ···················· | ···················· | ···················· | ···················· | ···················· |
| J | 1199 | hCoV-19/England/MILK-83C05E/2020\|EPI_ISL_557222\|20 | ···················· | ···················· | ···················· | ···················· | ···················· | ···················· |
| J | 1200 | hCoV-19/England/MILK-8134D9/2020\|EPI_ISL_557223\|20 | ···················· | ···················· | ···················· | ···················· | ···················· | ···················· |
| J | 1201 | hCoV-19/England/MILK-8009A3/2020\|EPI_ISL_557224\|20 | ···················· | ···················· | ···················· | ···················· | ···················· | ···················· |
| J | 1202 | hCoV-19/England/MILK-804F61/2020\|EPI_ISL_557226\|20 | ···················· | ···················· | ···················· | ···················· | ···················· | ···················· |
| J | 1203 | hCoV-19/England/MILK-87C1C8/2020\|EPI_ISL_557228\|20 | ···················· | ···················· | ···················· | ···················· | ···················· | ···················· |
| J | 1204 | hCoV-19/England/MILK-8249F4/2020\|EPI_ISL_557230\|20 | ···················· | ···················· | ···················· | ···················· | ···················· | ···················· |
| J | 1205 | hCoV-19/England/MILK-87A4B2/2020\|EPI_ISL_557231\|20 | ···················· | ·A·················· | ···················· | ···················· | ···················· | ···················· |
| J | 1206 | hCoV-19/England/MILK-80F591/2020\|EPI_ISL_557233\|20 | ···················· | ···················· | ···················· | ···················· | ···················· | ···················· |
| J | 1207 | hCoV-19/England/MILK-833312/2020\|EPI_ISL_557234\|20 | ···················· | ···················· | ···················· | ···················· | ···················· | ···················· |
| J | 1208 | hCoV-19/England/MILK-839121/2020\|EPI_ISL_557235\|20 | ···················· | ···················· | ···················· | ···················· | ···················· | ···················· |
| J | 1209 | hCoV-19/England/MILK-85A148/2020\|EPI_ISL_557236\|20 | ···················· | ···················· | ···················· | ···················· | ···················· | ···················· |
| J | 1210 | hCoV-19/England/MILK-82D4AF/2020\|EPI_ISL_557239\|20 | ···················· | ···················· | ···················· | ···················· | ···················· | ···················· |
| J | 1211 | hCoV-19/England/ALDP-5CB0C0/2020\|EPI_ISL_558527\|20 | ···················· | ···················· | ···················· | ···················· | ···················· | ···················· |
| J | 1212 | hCoV-19/England/ALDP-5CB50D/2020\|EPI_ISL_558528\|20 | ···················· | ···················· | ···················· | ···················· | ···················· | ···················· |
| J | 1213 | hCoV-19/England/ALDP-5C664B/2020\|EPI_ISL_558529\|20 | ···················· | ·············A······ | ···················· | ···················· | ···················· | ···················· |
| J | 1214 | hCoV-19/England/ALDP-5CFC40/2020\|EPI_ISL_558531\|20 | ···················· | ···················· | ···················· | ···················· | ···················· | ···················· |
| J | 1215 | hCoV-19/England/ALDP-5D2BA7/2020\|EPI_ISL_558532\|20 | ···················· | ···················· | ···················· | ···················· | ···················· | ···················· |
| J | 1216 | hCoV-19/England/ALDP-5D2749/2020\|EPI_ISL_558533\|20 | ···················· | ···················· | ···················· | ···················· | ···················· | ···················· |
| J | 1217 | hCoV-19/England/ALDP-5D0189/2020\|EPI_ISL_558534\|20 | ···················· | ···················· | ···················· | ···················· | ···················· | ···················· |
| J | 1218 | hCoV-19/England/ALDP-5D356C/2020\|EPI_ISL_558535\|20 | ···················· | ···················· | ···················· | ···················· | ···················· | ···················· |
| J | 1219 | hCoV-19/England/ALDP-5D347E/2020\|EPI_ISL_558536\|20 | ···················· | ···················· | ···················· | ···················· | ···················· | ···················· |
| J | 1220 | hCoV-19/England/ALDP-5CA9F3/2020\|EPI_ISL_558537\|20 | ···················· | ···················· | ···················· | ···················· | ···················· | ···················· |
| J | 1221 | hCoV-19/England/ALDP-5C99D6/2020\|EPI_ISL_558538\|20 | ···················· | ···················· | ···················· | ···················· | ···················· | ···················· |
| J | 1222 | hCoV-19/England/ALDP-5D19BD/2020\|EPI_ISL_558539\|20 | ···················· | ···················· | ···················· | ···················· | ···················· | ···················· |
| J | 1223 | hCoV-19/England/ALDP-5D3DEC/2020\|EPI_ISL_558540\|20 | ···················· | ···················· | ···················· | ···················· | ···················· | ···················· |
| J | 1224 | hCoV-19/England/ALDP-5D3960/2020\|EPI_ISL_558541\|20 | ···················· | ···················· | ···················· | ···················· | ···················· | ···················· |
| J | 1225 | hCoV-19/England/ALDP-5CD0DD/2020\|EPI_ISL_558542\|20 | ···················· | ···················· | ···················· | ···················· | ···················· | ···················· |
| J | 1226 | hCoV-19/England/ALDP-5D210F/2020\|EPI_ISL_558544\|20 | ···················· | ···················· | ···················· | ···················· | ···················· | ···················· |
| J | 1227 | hCoV-19/England/ALDP-5C87A0/2020\|EPI_ISL_558546\|20 | ···················· | ···················· | ···················· | ···················· | ···················· | ···················· |
| J | 1228 | hCoV-19/England/ALDP-5CEFFD/2020\|EPI_ISL_558547\|20 | ···················· | ···················· | ···················· | ···················· | ···················· | ···················· |
| J | 1229 | hCoV-19/England/ALDP-5C711C/2020\|EPI_ISL_558548\|20 | ···················· | ···················· | ···················· | ···················· | ···················· | ···················· |
| J | 1230 | hCoV-19/England/ALDP-5D5360/2020\|EPI_ISL_558549\|20 | ···················· | ···················· | ···················· | ···················· | ···················· | ···················· |
| J | 1231 | hCoV-19/England/ALDP-5D3C58/2020\|EPI_ISL_558550\|20 | ···················· | ···················· | ···················· | ···················· | ···················· | ···················· |
| J | 1232 | hCoV-19/England/ALDP-5C9D52/2020\|EPI_ISL_558551\|20 | ···················· | ···················· | ···················· | ···················· | ···················· | ···················· |
| J | 1233 | hCoV-19/England/ALDP-5CFFDE/2020\|EPI_ISL_558552\|20 | ···················· | ···················· | ···················· | ···················· | ···················· | ···················· |
| J | 1234 | hCoV-19/England/ALDP-5D41D0/2020\|EPI_ISL_558555\|20 | ···················· | ···················· | ···················· | ···················· | ···················· | ···················· |
| J | 1235 | hCoV-19/England/ALDP-5C7E7F/2020\|EPI_ISL_558556\|20 | ···················· | ···················· | ···················· | ···················· | ···················· | ···················· |
| J | 1236 | hCoV-19/England/ALDP-5C74E6/2020\|EPI_ISL_558557\|20 | ···················· | ···················· | ···················· | ···················· | ···················· | ···················· |
| J | 1237 | hCoV-19/England/ALDP-5CCCDA/2020\|EPI_ISL_558560\|20 | ···················· | ·A·················· | ···················· | ···················· | ···················· | ···················· |
| J | 1238 | hCoV-19/England/ALDP-5C9657/2020\|EPI_ISL_558562\|20 | ···················· | ···················· | ···················· | ···················· | ···················· | ···················· |
| J | 1239 | hCoV-19/England/ALDP-5C7C66/2020\|EPI_ISL_558563\|20 | ···················· | ···················· | ···················· | ···················· | ···················· | ···················· |
| J | 1240 | hCoV-19/England/ALDP-5D5157/2020\|EPI_ISL_558564\|20 | ···················· | ···················· | ···················· | ···················· | ···················· | ···················· |
| J | 1241 | hCoV-19/England/ALDP-5C7765/2020\|EPI_ISL_558566\|20 | ···················· | ···················· | ···················· | ···················· | ···················· | ···················· |
| J | 1242 | hCoV-19/England/ALDP-5D65A5/2020\|EPI_ISL_558568\|20 | ···················· | ·A·················· | ···················· | ···················· | ···················· | ···················· |
| J | 1243 | hCoV-19/England/ALDP-5D1BD5/2020\|EPI_ISL_558569\|20 | ···················· | ·A·················· | ···················· | ···················· | ···················· | ···················· |
| J | 1244 | hCoV-19/England/QEUH-9B0C7B/2020\|EPI_ISL_559858\|20 | ···················· | ···················· | ···················· | ···················· | ···················· | ···················· |
| J | 1245 | hCoV-19/England/QEUH-9B0CA8/2020\|EPI_ISL_559859\|20 | ···················· | ···················· | ···················· | ···················· | ···················· | ···················· |
| J | 1246 | hCoV-19/England/QEUH-9B0CD5/2020\|EPI_ISL_559860\|20 | ···················· | ···················· | ···················· | ···················· | ····T··············· | ···················· |
| J | 1247 | hCoV-19/England/QEUH-9B0D0F/2020\|EPI_ISL_559861\|20 | ···················· | ···················· | ···················· | ···················· | ···················· | ···················· |
| J | 1248 | hCoV-19/England/QEUH-9B0D1E/2020\|EPI_ISL_559862\|20 | ···················· | ···················· | ···················· | ···················· | ···················· | ···················· |
| J | 1249 | hCoV-19/England/QEUH-9B0D2D/2020\|EPI_ISL_559863\|20 | ···················· | ···················· | ···················· | ···················· | ····T··············· | ···················· |
| J | 1250 | hCoV-19/England/QEUH-9B0D3C/2020\|EPI_ISL_559864\|20 | ···················· | ···················· | ···················· | ···················· | ···················· | ···················· |
| J | 1251 | hCoV-19/England/QEUH-9B0D5A/2020\|EPI_ISL_559865\|20 | ···················· | ···················· | ···················· | ···················· | ···················· | ···················· |
| J | 1252 | hCoV-19/England/QEUH-9B0DF0/2020\|EPI_ISL_559866\|20 | ···················· | ···················· | ···················· | ···················· | ···················· | ···················· |
| J | 1253 | hCoV-19/England/QEUH-9B0E39/2020\|EPI_ISL_559867\|20 | ···················· | ···················· | ···················· | ···················· | ···················· | ···················· |
| J | 1254 | hCoV-19/England/QEUH-9B0F36/2020\|EPI_ISL_559868\|20 | ···················· | ···················· | ···················· | ···················· | ···················· | ···················· |
| J | 1255 | hCoV-19/England/QEUH-9B0F45/2020\|EPI_ISL_559869\|20 | ···················· | ···················· | ···················· | ···················· | ···················· | ···················· |

| | | | | | | | | |
|---|---|---|---|---|---|---|---|---|
| J | 1256 | hCoV-19/England/QEUH-9B0F54/2020\|EPI_ISL_559870\|20 | ···················· | ···················· | ···················· | ···················· | ···················· | ···················· |
| J | 1257 | hCoV-19/England/QEUH-9B0F63/2020\|EPI_ISL_559871\|20 | ···················· | ···················· | ···················· | ···················· | ···················· | ···················· |
| J | 1258 | hCoV-19/England/QEUH-9B0F81/2020\|EPI_ISL_559872\|20 | ···················· | ···················· | ···················· | ···················· | ···················· | ···················· |
| J | 1259 | hCoV-19/England/QEUH-9B3FBB/2020\|EPI_ISL_559873\|20 | ···················· | ···················· | ···················· | ···················· | ···················· | ···················· |
| J | 1260 | hCoV-19/England/QEUH-9B3FCA/2020\|EPI_ISL_559874\|20 | ···················· | ···················· | ···················· | ···················· | ···················· | ···················· |
| J | 1261 | hCoV-19/England/QEUH-9B3FD9/2020\|EPI_ISL_559875\|20 | ···················· | ···················· | ···················· | ···················· | ···················· | ···················· |
| J | 1262 | hCoV-19/England/QEUH-9B4011/2020\|EPI_ISL_559876\|20 | ···················· | ···················· | ···················· | ···················· | ···················· | ···················· |
| J | 1263 | hCoV-19/England/QEUH-9B405D/2020\|EPI_ISL_559877\|20 | ···················· | ···················· | ···················· | ···················· | ···················· | ···················· |
| J | 1264 | hCoV-19/England/QEUH-9B40F3/2020\|EPI_ISL_559878\|20 | ···················· | ···················· | ···················· | ···················· | ···················· | ···················· |
| J | 1265 | hCoV-19/England/QEUH-9B414B/2020\|EPI_ISL_559879\|20 | ···················· | ···················· | ···················· | ···················· | ···················· | ···················· |
| J | 1266 | hCoV-19/England/QEUH-9B41E1/2020\|EPI_ISL_559881\|20 | ·······T············ | ···················· | ···················· | ···················· | ···················· | ···················· |
| J | 1267 | hCoV-19/England/QEUH-9B422A/2020\|EPI_ISL_559882\|20 | ···················· | ·A·················· | ···················· | ···················· | ···················· | ···················· |
| J | 1268 | hCoV-19/England/QEUH-9B4266/2020\|EPI_ISL_559883\|20 | ···················· | ···················· | ···················· | ···················· | ···················· | ···················· |
| J | 1269 | hCoV-19/England/QEUH-9B4275/2020\|EPI_ISL_559884\|20 | ···················· | ···················· | ···················· | ···················· | ···················· | ···················· |
| J | 1270 | hCoV-19/England/QEUH-9B4284/2020\|EPI_ISL_559885\|20 | ···················· | ···················· | ···················· | ···················· | ···················· | ···················· |
| J | 1271 | hCoV-19/England/QEUH-9B42C0/2020\|EPI_ISL_559887\|20 | ···················· | ···················· | ···················· | ···················· | ···················· | ···················· |
| J | 1272 | hCoV-19/England/QEUH-9B43CD/2020\|EPI_ISL_559890\|20 | ···················· | ···················· | ···················· | ···················· | ···················· | ···················· |
| J | 1273 | hCoV-19/England/QEUH-9B4406/2020\|EPI_ISL_559891\|20 | ···················· | ···················· | ···················· | ···················· | ···················· | ···················· |
| J | 1274 | hCoV-19/England/QEUH-9B4433/2020\|EPI_ISL_559892\|20 | ···················· | ···················· | ···················· | ···················· | ···················· | ···················· |
| J | 1275 | hCoV-19/England/QEUH-9B447F/2020\|EPI_ISL_559893\|20 | ···················· | ···················· | ···················· | ···················· | ···················· | ···················· |
| J | 1276 | hCoV-19/England/QEUH-9B448E/2020\|EPI_ISL_559894\|20 | ···················· | ···················· | ···················· | ···················· | ···················· | ···················· |
| J | 1277 | hCoV-19/England/QEUH-9B44AC/2020\|EPI_ISL_559895\|20 | ···················· | ···················· | ···················· | ···················· | ·····T·············· | ···················· |
| J | 1278 | hCoV-19/England/QEUH-9B457C/2020\|EPI_ISL_559896\|20 | ···················· | ···················· | ···················· | ···················· | ···················· | ···················· |
| J | 1279 | hCoV-19/England/QEUH-9B4794/2020\|EPI_ISL_559897\|20 | ···················· | ···················· | ···················· | ···················· | ···················· | ···················· |
| J | 1280 | hCoV-19/England/SHEF-C22D7/2020\|EPI_ISL_559899\|202 | ···················· | ···················· | ···················· | ···················· | ···················· | ···················· |
| J | 1281 | hCoV-19/England/MILK-9AB341/2020\|EPI_ISL_551059\|20 | ···················· | ···················· | ···················· | ···················· | ···················· | ···················· |
| J | 1282 | hCoV-19/England/MILK-9AB183/2020\|EPI_ISL_551060\|20 | ···················· | ·A·················· | ···················· | ···················· | ···················· | ··A················· |
| J | 1283 | hCoV-19/England/MILK-9AB323/2020\|EPI_ISL_551061\|20 | ···················· | ·A·················· | ···················· | ···················· | ···················· | ··A················· |
| J | 1284 | hCoV-19/England/MILK-9AB4B7/2020\|EPI_ISL_551062\|20 | ···················· | ···················· | ···················· | ···················· | ···················· | ···················· |
| J | 1285 | hCoV-19/England/MILK-9A9C39/2020\|EPI_ISL_551064\|20 | ···················· | ···················· | ···················· | ···················· | ···················· | ···················· |
| J | 1286 | hCoV-19/England/MILK-9A9659/2020\|EPI_ISL_551065\|20 | ···················· | ···················· | ···················· | ···················· | ···················· | ···················· |
| J | 1287 | hCoV-19/England/MILK-9AB11A/2020\|EPI_ISL_551066\|20 | ···················· | ·A·················· | ···················· | ···················· | ···················· | ··A················· |
| J | 1288 | hCoV-19/England/MILK-9A9756/2020\|EPI_ISL_551067\|20 | ···················· | ···················· | ···················· | ···················· | ···················· | ···················· |
| J | 1289 | hCoV-19/England/MILK-9AB01D/2020\|EPI_ISL_551068\|20 | ···················· | ·A·················· | ···················· | ···················· | ···················· | ··A················· |
| J | 1290 | hCoV-19/England/MILK-9AB059/2020\|EPI_ISL_551069\|20 | ···················· | ···················· | ···················· | ···················· | ···················· | ···················· |
| J | 1291 | hCoV-19/England/MILK-9A9C48/2020\|EPI_ISL_551070\|20 | ···················· | ···················· | ···················· | ···················· | ···················· | ···················· |
| J | 1292 | hCoV-19/England/MILK-9AB402/2020\|EPI_ISL_551071\|20 | ···················· | ···················· | ···················· | ···················· | ···················· | ···················· |
| J | 1293 | hCoV-19/England/MILK-9A9185/2020\|EPI_ISL_551073\|20 | ···················· | ···················· | ···················· | ···················· | ···················· | ···················· |
| J | 1294 | hCoV-19/England/MILK-9A9EE8/2020\|EPI_ISL_551074\|20 | ···················· | ···················· | ···················· | ···················· | ···················· | ···················· |
| J | 1295 | hCoV-19/England/MILK-9AB1FC/2020\|EPI_ISL_551075\|20 | ···················· | ···················· | ···················· | ···················· | ···················· | ···················· |
| J | 1296 | hCoV-19/England/MILK-9AB156/2020\|EPI_ISL_551076\|20 | ···················· | ·A·················· | ···················· | ···················· | ···················· | ··A················· |
| J | 1297 | hCoV-19/England/MILK-9A9A5D/2020\|EPI_ISL_551077\|20 | ···················· | ···················· | ···················· | ···················· | ···················· | ···················· |
| J | 1298 | hCoV-19/England/MILK-9A93CB/2020\|EPI_ISL_551078\|20 | ···················· | ·A·················· | ···················· | ···················· | ···················· | ···················· |
| J | 1299 | hCoV-19/England/MILK-9A9176/2020\|EPI_ISL_551080\|20 | ···················· | ···················· | ···················· | ···················· | ···················· | ···················· |
| J | 1300 | hCoV-19/England/MILK-9A9E9D/2020\|EPI_ISL_551081\|20 | ···················· | ···················· | ···················· | ···················· | ···················· | ···················· |
| J | 1301 | hCoV-19/England/MILK-9A9BA5/2020\|EPI_ISL_551082\|20 | ···················· | ···················· | ···················· | ···················· | ···················· | ···················· |
| J | 1302 | hCoV-19/England/MILK-9AB1A1/2020\|EPI_ISL_551083\|20 | ···················· | ···················· | ···················· | ···················· | ···················· | ···················· |
| J | 1303 | hCoV-19/England/MILK-9AB208/2020\|EPI_ISL_551084\|20 | ···················· | ···················· | ···················· | ···················· | ···················· | ···················· |
| J | 1304 | hCoV-19/England/MILK-9A90B5/2020\|EPI_ISL_551085\|20 | ···················· | ···················· | ···················· | ···················· | ···················· | ···················· |
| J | 1305 | hCoV-19/England/MILK-9AB2CC/2020\|EPI_ISL_551086\|20 | ···················· | ···················· | ···················· | ···················· | ···T················ | ···················· |
| J | 1306 | hCoV-19/England/MILK-9A9431/2020\|EPI_ISL_551087\|20 | ···················· | ···················· | ···················· | ···················· | ·····T·············· | ···················· |
| J | 1307 | hCoV-19/England/MILK-9A9DBE/2020\|EPI_ISL_551088\|20 | ···················· | ···················· | ···················· | ···················· | ···················· | ···················· |
| J | 1308 | hCoV-19/England/MILK-9A93DA/2020\|EPI_ISL_551089\|20 | ···················· | ···················· | ···················· | ···················· | ···················· | ···················· |
| J | 1309 | hCoV-19/England/MILK-9AB280/2020\|EPI_ISL_551090\|20 | ···················· | ···················· | ···················· | ···················· | ···················· | ···················· |
| J | 1310 | hCoV-19/England/MILK-9AB314/2020\|EPI_ISL_551093\|20 | ···················· | ···················· | ···················· | ···················· | ···················· | ···················· |
| J | 1311 | hCoV-19/England/MILK-9A9CC0/2020\|EPI_ISL_551094\|20 | ···················· | ···················· | ···················· | ···················· | ···················· | ···················· |
| J | 1312 | hCoV-19/England/MILK-9AB271/2020\|EPI_ISL_551095\|20 | ···················· | ···················· | ···················· | ···················· | ···················· | ···················· |
| J | 1313 | hCoV-19/England/MILK-9A93E9/2020\|EPI_ISL_551096\|20 | ···················· | ···················· | ···················· | ···················· | ·····T·············· | ···················· |
| J | 1314 | hCoV-19/England/MILK-9AB50F/2020\|EPI_ISL_551097\|20 | ···················· | ···················· | ···················· | ···················· | ···················· | ···················· |
| J | 1315 | hCoV-19/England/MILK-9A9765/2020\|EPI_ISL_551098\|20 | ···················· | ·A······A··········· | ···················· | ···················· | ···················· | ···················· |
| J | 1316 | hCoV-19/England/MILK-9AB4D5/2020\|EPI_ISL_551099\|20 | ···················· | ···················· | ···················· | ···················· | ···················· | ···················· |
| J | 1317 | hCoV-19/England/MILK-9A8F7D/2020\|EPI_ISL_551100\|20 | ···················· | ···················· | ···················· | ···················· | ···················· | ···················· |
| J | 1318 | hCoV-19/England/MILK-9A9158/2020\|EPI_ISL_551101\|20 | ···················· | ···················· | ···················· | ···················· | ···················· | ···················· |
| J | 1319 | hCoV-19/England/MILK-9AAF6C/2020\|EPI_ISL_551102\|20 | ···················· | ···················· | ···················· | ···················· | ···················· | ···················· |
| J | 1320 | hCoV-19/England/MILK-9AB0C2/2020\|EPI_ISL_551103\|20 | ···················· | ···················· | ···················· | ···················· | ···················· | ···················· |
| J | 1321 | hCoV-19/England/MILK-9A9422/2020\|EPI_ISL_551104\|20 | ···················· | ···················· | ···················· | ···················· | ·····T·············· | ···················· |
| J | 1322 | hCoV-19/England/MILK-9AB226/2020\|EPI_ISL_551107\|20 | ···················· | ···················· | ···················· | ···················· | ···················· | ···················· |
| J | 1323 | hCoV-19/England/MILK-9A91B2/2020\|EPI_ISL_551109\|20 | ···················· | ···················· | ···················· | ···················· | ···················· | ···················· |
| J | 1324 | hCoV-19/England/MILK-9A97CF/2020\|EPI_ISL_551110\|20 | ···················· | ···················· | ···················· | ···················· | ···················· | ···················· |
| J | 1325 | hCoV-19/England/MILK-9A95C5/2020\|EPI_ISL_551111\|20 | ···················· | ···················· | ···················· | ···················· | ···················· | ···················· |
| J | 1326 | hCoV-19/England/MILK-9A9219/2020\|EPI_ISL_551112\|20 | ···················· | ···················· | ···················· | ···················· | ···················· | ···················· |
| J | 1327 | hCoV-19/England/MILK-9AB39C/2020\|EPI_ISL_551113\|20 | ···················· | ···················· | ···················· | ···················· | ···················· | ···················· |
| J | 1328 | hCoV-19/England/MILK-9A8FAA/2020\|EPI_ISL_551115\|20 | ···················· | ···················· | ···················· | ···················· | ···················· | ···················· |
| J | 1329 | hCoV-19/England/MILK-9AB244/2020\|EPI_ISL_551116\|20 | ···················· | ···················· | ···················· | ···················· | ···················· | ···················· |
| J | 1330 | hCoV-19/England/MILK-9A9AE4/2020\|EPI_ISL_551117\|20 | ···················· | ···················· | ···················· | ···················· | ···················· | ···················· |
| J | 1331 | hCoV-19/England/MILK-9A9149/2020\|EPI_ISL_551118\|20 | ···················· | ···················· | ···················· | ···················· | ···················· | ···················· |
| J | 1332 | hCoV-19/England/MILK-9A9AB7/2020\|EPI_ISL_551119\|20 | ···················· | ···················· | ···················· | ···················· | ···················· | ···················· |
| J | 1333 | hCoV-19/England/MILK-9AB192/2020\|EPI_ISL_551120\|20 | ···················· | ···················· | ···················· | ···················· | ···················· | ···················· |
| J | 1334 | hCoV-19/England/MILK-9A96C2/2020\|EPI_ISL_551121\|20 | ···················· | ···················· | ···················· | ···················· | ···················· | ···················· |
| J | 1335 | hCoV-19/England/MILK-9A963B/2020\|EPI_ISL_551122\|20 | ···················· | ···················· | ···················· | ···················· | ···················· | ···················· |

J 1336 hCoV-19/England/MILK-9AB2EA/2020|EPI_ISL_551131|20 ···················· ···················· ····················· ····················· ···················· ····················
J 1337 hCoV-19/England/MILK-9A93AD/2020|EPI_ISL_551132|20 ···················· ···················· ····················· ····················· ···················· ····················
J 1338 hCoV-19/England/MILK-9A9343/2020|EPI_ISL_551133|20 ···················· ···················· ····················· ····················· ···················· ····················
J 1339 hCoV-19/England/MILK-994A37/2020|EPI_ISL_552389|20 ···················· ···················· ····················· ····················· ···················· ····················
J 1340 hCoV-19/England/MILK-995140/2020|EPI_ISL_552393|20 ···················· ···················· ····················· ····················· ···················· ····················
J 1341 hCoV-19/England/MILK-993CC9/2020|EPI_ISL_552394|20 ···················· ···················· ····················· ····················· ···················· ····················
J 1342 hCoV-19/England/MILK-9945CD/2020|EPI_ISL_552395|20 ···················· ···················· ····················· ····················· ···················· ····················
J 1343 hCoV-19/England/MILK-9953E0/2020|EPI_ISL_552396|20 ···················· ·A·················· ····················· ····················· ···················· ····················
J 1344 hCoV-19/England/MILK-994E86/2020|EPI_ISL_552397|20 ···················· ·A·················· ····················· ····················· ···················· ···A················
J 1345 hCoV-19/England/MILK-991C9E/2020|EPI_ISL_552398|20 ···················· ···················· ····················· ····················· ···················· ····················
J 1346 hCoV-19/England/MILK-99477C/2020|EPI_ISL_552422|20 ···················· ···················· ····················· ····················· ···················· ····················
J 1347 hCoV-19/England/MILK-994B16/2020|EPI_ISL_552423|20 ···················· ···A················ ····················· ····················· ···················· ····················
J 1348 hCoV-19/England/MILK-994FDE/2020|EPI_ISL_552426|20 ···················· ···················· ····················· ····················· ···················· ····················
J 1349 hCoV-19/England/MILK-991D9B/2020|EPI_ISL_552427|20 ···················· ···················· ····················· ····················· ···················· ····················
J 1350 hCoV-19/England/MILK-993A92/2020|EPI_ISL_552428|20 ···················· ···················· ····················· ····················· ···················· ····················
J 1351 hCoV-19/England/MILK-994A28/2020|EPI_ISL_552429|20 ···················· ···················· ····················· ····················· ···················· ····················
J 1352 hCoV-19/England/MILK-993D4E/2020|EPI_ISL_552430|20 ···················· ·A·················· ····················· ····················· ···················· ···A················
J 1353 hCoV-19/England/MILK-994C40/2020|EPI_ISL_552431|20 ···················· ···················· ····················· ····················· ···················· ····················
J 1354 hCoV-19/England/MILK-994C9B/2020|EPI_ISL_552432|20 ···················· ···················· ····················· ····················· ······T············· ····················
J 1355 hCoV-19/England/MILK-995517/2020|EPI_ISL_552433|20 ···················· ···················· ····················· ····················· ···················· ····················
J 1356 hCoV-19/England/MILK-994EC2/2020|EPI_ISL_552436|20 ···················· ···················· ····················· ····················· ···················· ····················
J 1357 hCoV-19/England/MILK-9951F5/2020|EPI_ISL_552437|20 ········T··········· ···················· ····················· ····················· ·Y·················· ····················
J 1358 hCoV-19/England/MILK-994E68/2020|EPI_ISL_552438|20 ···················· ···················· ····················· ····················· ···················· ····················
J 1359 hCoV-19/England/MILK-994888/2020|EPI_ISL_552439|20 ···················· ·A·················· ····················· ····················· ···················· ···A················
J 1360 hCoV-19/England/MILK-993A74/2020|EPI_ISL_552440|20 ···················· ···················· ····················· ····················· ···················· ····················
J 1361 hCoV-19/Wales/MILK-9950E9/2020|EPI_ISL_552441|2020 ···················· ···················· ····················· ····················· ···················· ····················
J 1362 hCoV-19/England/MILK-994E1D/2020|EPI_ISL_552442|20 ···················· ···················· ····················· ····················· ···················· ····················
J 1363 hCoV-19/England/MILK-993A47/2020|EPI_ISL_552443|20 ···················· ···················· ····················· ····················· ···················· ····················
J 1364 hCoV-19/England/MILK-975850/2020|EPI_ISL_552444|20 ···················· ···················· ····················· ····················· ···················· ····················
J 1365 hCoV-19/England/MILK-9751FB/2020|EPI_ISL_552445|20 ···················· ···················· ····················· ····················· ···················· ····················
J 1366 hCoV-19/England/ALDP-982999/2020|EPI_ISL_552446|20 ···················· ···················· ····················· ····················· ···················· ····················
J 1367 hCoV-19/England/MILK-9750FE/2020|EPI_ISL_552448|20 ···················· ···················· ····················· ····················· ···················· ····················
J 1368 hCoV-19/England/ALDP-9829C6/2020|EPI_ISL_552449|20 ···················· ···················· ····················· ····················· ···················· ····················
J 1369 hCoV-19/England/MILK-975683/2020|EPI_ISL_552450|20 ···················· ···················· ····················· ····················· ···················· ····················
J 1370 hCoV-19/England/MILK-975085/2020|EPI_ISL_552451|20 ···················· ···················· ····················· ····················· ···················· ····················
J 1371 hCoV-19/England/MILK-974D70/2020|EPI_ISL_552452|20 ···················· ···················· ····················· ····················· ···················· ····················
J 1372 hCoV-19/England/MILK-975C72/2020|EPI_ISL_552453|20 ···················· ···················· ····················· ····················· ···················· ····················
J 1373 hCoV-19/England/MILK-974FC5/2020|EPI_ISL_552454|20 ···················· ···················· ····················· ····················· ···················· ····················
J 1374 hCoV-19/England/MILK-975261/2020|EPI_ISL_552462|20 ···················· ·A·················· ····················· ····················· ···················· ···A················
J 1375 hCoV-19/England/ALDP-982A5A/2020|EPI_ISL_552463|20 ···················· ···················· ····················· ····················· ···················· ····················
J 1376 hCoV-19/England/ALDP-982C09/2020|EPI_ISL_552464|20 ···················· ···················· ····················· ····················· ···················· ····················
J 1377 hCoV-19/England/MILK-9752DA/2020|EPI_ISL_552465|20 ···················· ···················· ····················· ····················· ···················· ····················
J 1378 hCoV-19/England/ALDP-9540C8/2020|EPI_ISL_553755|20 ···················· ···················· ····················· ····················· ···················· ····················
J 1379 hCoV-19/England/ALDP-9537A6/2020|EPI_ISL_553756|20 ···················· ···················· ····················· ····················· ···················· ····················
J 1380 hCoV-19/England/ALDP-9543ED/2020|EPI_ISL_553760|20 ···················· ···················· ····················· ····················· ···················· ····················
J 1381 hCoV-19/England/ALDP-953524/2020|EPI_ISL_553761|20 ···················· ···················· ····················· ····················· ···················· ····················
J 1382 hCoV-19/England/ALDP-953BAA/2020|EPI_ISL_553762|20 ···················· ···················· ····················· ····················· ···················· ····················
J 1383 hCoV-19/England/ALDP-954AF7/2020|EPI_ISL_553763|20 ···················· ···················· ····················· ····················· ···················· ····················
J 1384 hCoV-19/England/ALDP-953973/2020|EPI_ISL_553764|20 ···················· ···················· ····················· ····················· ···················· ····················
J 1385 hCoV-19/England/ALDP-953955/2020|EPI_ISL_553765|20 ···················· ···················· ····················· ····················· ···················· ····················
J 1386 hCoV-19/England/ALDP-953C2F/2020|EPI_ISL_553766|20 ···················· ···················· ····················· ····················· ···················· ····················
J 1387 hCoV-19/England/ALDP-9542D1/2020|EPI_ISL_553767|20 ···················· ···················· ····················· ····················· ···················· ····················
J 1388 hCoV-19/England/ALDP-953BB9/2020|EPI_ISL_553768|20 ···················· ···················· ····················· ····················· ···················· ····················
J 1389 hCoV-19/England/ALDP-9541E3/2020|EPI_ISL_553769|20 ···················· ···················· ····················· ····················· ···················· ····················
J 1390 hCoV-19/Wales/ALDP-9541B6/2020|EPI_ISL_553770|2020 ···················· ···················· ····················· ····················· ···················· ····················
J 1391 hCoV-19/England/ALDP-953612/2020|EPI_ISL_553771|20 ···················· ···················· ····················· ····················· ···················· ····················
J 1392 hCoV-19/England/ALDP-95357F/2020|EPI_ISL_553772|20 ···················· ·A·················· ····················· ····················· ···················· ····················
J 1393 hCoV-19/England/ALDP-953B31/2020|EPI_ISL_553773|20 ···················· ···················· ····················· ····················· ···················· ····················
J 1394 hCoV-19/England/ALDP-953542/2020|EPI_ISL_553775|20 ···················· ···················· ····················· ····················· ···················· ····················
J 1395 hCoV-19/England/ALDP-954620/2020|EPI_ISL_553776|20 ···················· ···················· ····················· ····················· ···················· ····················
J 1396 hCoV-19/England/ALDP-953B6E/2020|EPI_ISL_553777|20 ···················· ···················· ····················· ····················· ···················· ····················
J 1397 hCoV-19/England/ALDP-953551/2020|EPI_ISL_553779|20 ···················· ···················· ····················· ····················· ···················· ····················
J 1398 hCoV-19/England/ALDP-95412F/2020|EPI_ISL_553781|20 ···················· ···················· ····················· ····················· ···················· ····················
J 1399 hCoV-19/England/ALDP-953621/2020|EPI_ISL_553782|20 ···················· ···················· ····················· ····················· ···················· ····················
J 1400 hCoV-19/England/ALDP-953964/2020|EPI_ISL_553783|20 ···················· ···················· ····················· ····················· ···················· ····················
J 1401 hCoV-19/England/ALDP-9538A3/2020|EPI_ISL_553785|20 ···················· ···················· ····················· ····················· ···················· ····················
J 1402 hCoV-19/England/ALDP-9538EF/2020|EPI_ISL_553791|20 ···················· ···················· ····················· ····················· ···················· ····················
J 1403 hCoV-19/England/ALDP-954444/2020|EPI_ISL_553793|20 ···················· ···················· ····················· ····················· ···················· ····················
J 1404 hCoV-19/England/ALDP-9539A0/2020|EPI_ISL_553795|20 ···················· ···················· ····················· ····················· ···················· ····················
J 1405 hCoV-19/England/ALDP-954198/2020|EPI_ISL_553796|20 ···················· ···················· ····················· ····················· ···················· ····················
J 1406 hCoV-19/England/MILK-82788B/2020|EPI_ISL_557240|20 ···················· ···················· ····················· ····················· ···················· ····················
J 1407 hCoV-19/England/MILK-87B6F7/2020|EPI_ISL_557241|20 ···················· ···················· ····················· ····················· ···················· ····················
J 1408 hCoV-19/England/MILK-820C4A/2020|EPI_ISL_557242|20 ···················· ···················· ····················· ····················· ···················· ····················
J 1409 hCoV-19/England/MILK-8731C1/2020|EPI_ISL_557244|20 ···················· ···················· ····················· ····················· ···················· ····················
J 1410 hCoV-19/England/MILK-82D937/2020|EPI_ISL_557246|20 ···················· ···················· ····················· ····················· ···················· ····················
J 1411 hCoV-19/England/MILK-845650/2020|EPI_ISL_557247|20 ···················· ···················· ····················· ····················· ···················· ····················
J 1412 hCoV-19/England/MILK-83CAE7/2020|EPI_ISL_557248|20 ···················· ·A·················· ····················· ····················· ···················· ····················
J 1413 hCoV-19/England/MILK-830360/2020|EPI_ISL_557251|20 ···················· ···················· ····················· ····················· ···················· ····················
J 1414 hCoV-19/England/MILK-84AE35/2020|EPI_ISL_557252|20 ···················· ···················· ····················· ····················· ···················· ····················
J 1415 hCoV-19/England/MILK-854278/2020|EPI_ISL_557253|20 ···················· ···················· ····················· ····················· ···················· ····················

```
J 1416 hCoV-19/England/MILK-879671/2020|EPI_ISL_557256|20 ···················· ···················· ···················· ···················· ···················· ····················
J 1417 hCoV-19/England/MILK-8054CB/2020|EPI_ISL_557257|20 ···················· ···················· ···················· ···················· ···················· ····················
J 1418 hCoV-19/England/MILK-819066/2020|EPI_ISL_557258|20 ···················· ···················· ···················· ···················· ···················· ····················
J 1419 hCoV-19/England/MILK-83DCA4/2020|EPI_ISL_557259|20 ···················· ···················· ···················· ···················· ···················· ····················
J 1420 hCoV-19/England/MILK-815413/2020|EPI_ISL_557260|20 ···················· ·A·················· ···················· ···················· ···················· ····················
J 1421 hCoV-19/England/MILK-8461E5/2020|EPI_ISL_557261|20 ···················· ···················· ···················· ···················· ···················· ····················
J 1422 hCoV-19/England/MILK-8328F6/2020|EPI_ISL_557263|20 ···················· ···················· ···················· ···················· ···················· ····················
J 1423 hCoV-19/England/MILK-817B76/2020|EPI_ISL_557264|20 ···················· ···················· ···················· ···················· ···················· ····················
J 1424 hCoV-19/England/MILK-809C29/2020|EPI_ISL_557266|20 ···················· ···················· ···········T········ ···················· ···················· ····················
J 1425 hCoV-19/England/MILK-83551A/2020|EPI_ISL_557267|20 ···················· ···················· ···················· ···················· ···················· ····················
J 1426 hCoV-19/England/MILK-832D60/2020|EPI_ISL_557268|20 ···················· ······A············· ···················· ···················· ···················· ····················
J 1427 hCoV-19/England/MILK-849DDF/2020|EPI_ISL_557270|20 ···················· ···················· ···················· ···················· ···················· ····················
J 1428 hCoV-19/England/MILK-81E3A4/2020|EPI_ISL_557273|20 ···················· ·A·················· ···················· ···················· ···················· ···A················
J 1429 hCoV-19/England/MILK-82264A/2020|EPI_ISL_557275|20 ···················· ·A·················· ···················· ···················· ···················· ····················
J 1430 hCoV-19/England/MILK-820819/2020|EPI_ISL_557276|20 ···················· ···················· ···················· ···················· ···················· ····················
J 1431 hCoV-19/England/MILK-87BC04/2020|EPI_ISL_557277|20 ···················· ···················· ···················· ···················· ···················· ····················
J 1432 hCoV-19/England/MILK-83CA8D/2020|EPI_ISL_557278|20 ···················· ·A·················· ···················· ···················· ···················· ····················
J 1433 hCoV-19/England/MILK-85465E/2020|EPI_ISL_557279|20 ···················· ···················· ···················· ···················· ···················· ····················
J 1434 hCoV-19/England/ALDP-5C7B78/2020|EPI_ISL_558501|20 ···················· ···················· ···················· ···················· ···················· ····················
J 1435 hCoV-19/England/ALDP-5D31A4/2020|EPI_ISL_558502|20 ···················· ···················· ···················· ···················· ···················· ····················
J 1436 hCoV-19/England/ALDP-5D271C/2020|EPI_ISL_558503|20 ···················· ···················· ···················· ···················· ···················· ····················
J 1437 hCoV-19/England/ALDP-5C961B/2020|EPI_ISL_558504|20 ···················· ···················· ···················· ···················· ···················· ····················
J 1438 hCoV-19/England/ALDP-5D0BB8/2020|EPI_ISL_558505|20 ···················· ···················· ···················· ···················· ···················· ····················
J 1439 hCoV-19/England/ALDP-5CDC24/2020|EPI_ISL_558506|20 ···················· ···················· ···················· ···················· ···················· ····················
J 1440 hCoV-19/England/ALDP-5D0A60/2020|EPI_ISL_558507|20 ···················· ···················· ···················· ···················· ···················· ····················
J 1441 hCoV-19/England/ALDP-5CE485/2020|EPI_ISL_558508|20 ···················· ···················· ···················· ···················· ···················· ····················
J 1442 hCoV-19/England/ALDP-5D58DA/2020|EPI_ISL_558510|20 ···················· ···················· ···················· ···················· ···················· ····················
J 1443 hCoV-19/England/ALDP-5D8C53/2020|EPI_ISL_558511|20 ···················· ···················· ···················· ···················· ···················· ····················
J 1444 hCoV-19/England/ALDP-5C5819/2020|EPI_ISL_558512|20 ···················· ···················· ···················· ···················· ···················· ····················
J 1445 hCoV-19/England/ALDP-5D7B84/2020|EPI_ISL_558513|20 ···················· ···················· ···················· ···················· ···················· ····················
J 1446 hCoV-19/England/ALDP-5D9AB2/2020|EPI_ISL_558514|20 ···················· ···················· ···················· ···················· ···················· ····················
J 1447 hCoV-19/England/ALDP-5CD1BC/2020|EPI_ISL_558515|20 ···················· ···················· ···················· ···················· ···················· ····················
J 1448 hCoV-19/England/ALDP-5D8AE0/2020|EPI_ISL_558516|20 ···················· ···················· ···················· ···················· ···················· ····················
J 1449 hCoV-19/England/ALDP-5C4FBB/2020|EPI_ISL_558517|20 ········T··········· ···················· ···················· ···················· ···················· ····················
J 1450 hCoV-19/England/ALDP-5CAAA5/2020|EPI_ISL_558518|20 ···················· ···················· ···················· ···················· ···················· ····················
J 1451 hCoV-19/England/ALDP-5D9D13/2020|EPI_ISL_558519|20 ···················· ···················· ···················· ···················· ···················· ····················
J 1452 hCoV-19/England/ALDP-5C701F/2020|EPI_ISL_558520|20 ···················· ···················· ···················· ···················· ···················· ····················
J 1453 hCoV-19/England/ALDP-5D1F15/2020|EPI_ISL_558521|20 ···················· ···················· ···················· ···················· ···················· ····················
J 1454 hCoV-19/England/ALDP-5D4E9D/2020|EPI_ISL_558522|20 ···················· ···················· ···················· ···················· ···················· ····················
J 1455 hCoV-19/England/ALDP-5C7F5E/2020|EPI_ISL_558523|20 ···················· ···················· ···················· ···················· ···················· ····················
J 1456 hCoV-19/England/ALDP-5D1FAC/2020|EPI_ISL_558525|20 ···················· ·A·················· ···················· ···················· ···················· ····················
J 1457 hCoV-19/England/ALDP-5CEF57/2020|EPI_ISL_558526|20 ···················· ···················· ···················· ···················· ···················· ····················
J 1458 hCoV-19/England/ALDP-5C78EA/2020|EPI_ISL_558559|20 ···················· ···················· ···················· ···················· ···················· ····················
J 1459 hCoV-19/England/ALDP-5D18ED/2020|EPI_ISL_558570|20 ···················· ·A·················· ···················· ···················· ···················· ····················
J 1460 hCoV-19/England/ALDP-5D5C1A/2020|EPI_ISL_558571|20 ···················· ···················· ···················· ···················· ···················· ····················
J 1461 hCoV-19/England/ALDP-5D87DA/2020|EPI_ISL_558573|20 ···················· ···················· ···················· ···················· ···················· ····················
J 1462 hCoV-19/England/ALDP-5D4D72/2020|EPI_ISL_558575|20 ···················· ···················· ···················· ···················· ···················· ····················
J 1463 hCoV-19/England/ALDP-5D9557/2020|EPI_ISL_558576|20 ···················· ···················· ···················· ···················· ···················· ····················
J 1464 hCoV-19/England/ALDP-5D4EAC/2020|EPI_ISL_558579|20 ···················· ···················· ···················· ···················· ···················· ····················
J 1465 hCoV-19/England/ALDP-5C9790/2020|EPI_ISL_558580|20 ···················· ···················· ···················· ···················· ···················· ····················
J 1466 hCoV-19/England/ALDP-5CB45B/2020|EPI_ISL_558581|20 ···················· ···················· ···················· ···················· ···················· ····················
J 1467 hCoV-19/England/ALDP-5CE8D4/2020|EPI_ISL_558582|20 ···················· ·A·················· ···················· ···················· ···················· ····················
J 1468 hCoV-19/England/ALDP-5D67EB/2020|EPI_ISL_558583|20 ···················· ·A·················· ···················· ···················· ···················· ····················
J 1469 hCoV-19/England/ALDP-5CC8B8/2020|EPI_ISL_558584|20 ···················· ···················· ···················· ···················· ···················· ····················
J 1470 hCoV-19/England/ALDP-5C5BB6/2020|EPI_ISL_558585|20 ···················· ···················· ···················· ···················· ···················· ····················
J 1471 hCoV-19/Wales/ALDP-5C49BD/2020|EPI_ISL_558587|2020 ···················· ···················· ···················· ···················· ···················· ····················
J 1472 hCoV-19/England/ALDP-5C606B/2020|EPI_ISL_558589|20 ···················· ···················· ···················· ···················· ···················· ····················
J 1473 hCoV-19/England/ALDP-5D2390/2020|EPI_ISL_558590|20 ···················· ···················· ···················· ···················· ···················· ····················
J 1474 hCoV-19/England/ALDP-5D5EC9/2020|EPI_ISL_558591|20 ···················· ···················· ···················· ···················· ···················· ····················
J 1475 hCoV-19/England/ALDP-5CF0BD/2020|EPI_ISL_558593|20 ···················· ···················· ···················· ···················· ···················· ····················
J 1476 hCoV-19/England/ALDP-5D1C69/2020|EPI_ISL_558595|20 ···················· ···················· ···················· ···················· ···················· ····················
J 1477 hCoV-19/England/ALDP-5D3195/2020|EPI_ISL_558596|20 ···················· ···················· ···················· ···················· ···················· ····················
J 1478 hCoV-19/England/ALDP-5D4DAF/2020|EPI_ISL_558597|20 ···················· ···················· ···················· ···················· ···················· ····················
J 1479 hCoV-19/England/ALDP-5CC87C/2020|EPI_ISL_558598|20 ···················· ···················· ···················· ···················· ···················· ····················
J 1480 hCoV-19/England/ALDP-5D33CC/2020|EPI_ISL_558599|20 ···················· ···················· ···················· ···················· ···················· ····················
J 1481 hCoV-19/England/QEUH-9ACBC0/2020|EPI_ISL_559800|20 ···················· ···················· ···················· ···················· ···················· ····················
J 1482 hCoV-19/England/QEUH-9ACBEE/2020|EPI_ISL_559801|20 ···················· ···················· ···················· ···················· ···················· ····················
J 1483 hCoV-19/England/QEUH-9ACC81/2020|EPI_ISL_559802|20 ···················· ·A·················· ···················· ···················· ···················· ····················
J 1484 hCoV-19/England/QEUH-9ACCAF/2020|EPI_ISL_559803|20 ···················· ···················· ···················· ···················· ···················· ····················
J 1485 hCoV-19/England/QEUH-9ACCCD/2020|EPI_ISL_559804|20 ···················· ···················· ···················· ···················· ······T············· ····················
J 1486 hCoV-19/England/QEUH-9ACCFA/2020|EPI_ISL_559805|20 ···················· ···················· ···················· ···················· ···················· ····················
J 1487 hCoV-19/England/QEUH-9ACD06/2020|EPI_ISL_559806|20 ···················· ·A·················· ···················· ···················· ···················· ····················
J 1488 hCoV-19/England/QEUH-9ACD24/2020|EPI_ISL_559807|20 ···················· ·A·················· ···················· ···················· ···················· ····················
J 1489 hCoV-19/England/QEUH-9ACD60/2020|EPI_ISL_559808|20 ···················· ···················· ···················· ···················· ···················· ····················
J 1490 hCoV-19/England/QEUH-9ACE03/2020|EPI_ISL_559809|20 ···················· ·A·················· ···················· ···················· ···················· ····················
J 1491 hCoV-19/England/QEUH-9ACE21/2020|EPI_ISL_559810|20 ···················· ···················· ···················· ···················· ···················· ····················
J 1492 hCoV-19/England/QEUH-9ACE5E/2020|EPI_ISL_559811|20 ···················· ···················· ···················· ···················· ···················· ····················
J 1493 hCoV-19/England/QEUH-9ACE7C/2020|EPI_ISL_559812|20 ···················· ···················· ···················· ···················· ···················· ····················
J 1494 hCoV-19/England/QEUH-9ACE8B/2020|EPI_ISL_559813|20 ···················· ···················· ···················· ···················· ···················· ····················
J 1495 hCoV-19/England/QEUH-9ACE9A/2020|EPI_ISL_559814|20 ···················· ·A·······A·········· ···················· ···················· ···················· ····················
```

| | | | | | | | |
|---|---|---|---|---|---|---|---|
| J | 1496 hCoV-19/England/QEUH-9ACF00/2020\|EPI_ISL_559815\|20 | ···················· | ···················· | ···················· | ···················· | ···················· | ···················· |
| J | 1497 hCoV-19/England/QEUH-9ACF79/2020\|EPI_ISL_559816\|20 | ···················· | ···················· | ···················· | ···················· | ···················· | ···················· |
| J | 1498 hCoV-19/England/QEUH-9AD093/2020\|EPI_ISL_559817\|20 | ···················· | ·A·················· | ···················· | ···················· | ···················· | ···A················ |
| J | 1499 hCoV-19/England/QEUH-9AD0DF/2020\|EPI_ISL_559818\|20 | ···················· | ···················· | ···················· | ···················· | ···················· | ···················· |
| J | 1500 hCoV-19/England/QEUH-9AD172/2020\|EPI_ISL_559819\|20 | ···················· | ···················· | ···················· | ···················· | ····T··············· | ···················· |
| J | 1501 hCoV-19/England/QEUH-9AD1BE/2020\|EPI_ISL_559820\|20 | ···················· | ···················· | ···················· | ···················· | ···················· | ···················· |
| J | 1502 hCoV-19/England/QEUH-9AD224/2020\|EPI_ISL_559821\|20 | ···················· | ···················· | ···················· | ···················· | ···················· | ···················· |
| J | 1503 hCoV-19/England/QEUH-9AD242/2020\|EPI_ISL_559822\|20 | ···················· | ···················· | ···················· | ···················· | ···················· | ···················· |
| J | 1504 hCoV-19/England/QEUH-9AD27F/2020\|EPI_ISL_559823\|20 | ···················· | ···················· | ···················· | ···················· | ···················· | ···················· |
| J | 1505 hCoV-19/England/QEUH-9AD2E8/2020\|EPI_ISL_559824\|20 | ···················· | ·A·················· | ···················· | ···················· | ···················· | ···················· |
| J | 1506 hCoV-19/England/QEUH-9AF1DA/2020\|EPI_ISL_559826\|20 | ···················· | ·A·················· | ···················· | ···················· | ···················· | ···················· |
| J | 1507 hCoV-19/England/QEUH-9AF1F8/2020\|EPI_ISL_559827\|20 | ···················· | ···················· | ···················· | ···················· | ···················· | ···················· |
| J | 1508 hCoV-19/England/QEUH-9AF204/2020\|EPI_ISL_559828\|20 | ···················· | ···················· | ···················· | ···················· | ····T··············· | ···················· |
| J | 1509 hCoV-19/England/QEUH-9AF222/2020\|EPI_ISL_559829\|20 | ···················· | ···················· | ···················· | ···················· | ···················· | ···················· |
| J | 1510 hCoV-19/England/QEUH-9AF29B/2020\|EPI_ISL_559830\|20 | ···················· | ···················· | ···················· | ···················· | ···················· | ···················· |
| J | 1511 hCoV-19/England/QEUH-9AF2AA/2020\|EPI_ISL_559831\|20 | ···················· | ···················· | ···················· | ···················· | ···················· | ···················· |
| J | 1512 hCoV-19/England/QEUH-9AF2B9/2020\|EPI_ISL_559832\|20 | ···················· | ·A·················· | ···················· | ···················· | ···················· | ···················· |
| J | 1513 hCoV-19/England/QEUH-9AF2E6/2020\|EPI_ISL_559834\|20 | ···················· | ···················· | ···················· | ···················· | ···················· | ···················· |
| J | 1514 hCoV-19/England/QEUH-9AF301/2020\|EPI_ISL_559835\|20 | ···················· | ···················· | ···················· | ···················· | ···················· | ···················· |
| J | 1515 hCoV-19/England/QEUH-9AF33E/2020\|EPI_ISL_559836\|20 | ···················· | ···················· | ···················· | ···················· | ···················· | ···················· |
| J | 1516 hCoV-19/England/QEUH-9AF35C/2020\|EPI_ISL_559837\|20 | ···················· | ···················· | ···················· | ···················· | ···················· | ···················· |
| J | 1517 hCoV-19/England/QEUH-9AF36B/2020\|EPI_ISL_559838\|20 | ···················· | ···················· | ···················· | ···················· | ···················· | ···················· |
| J | 1518 hCoV-19/England/QEUH-9AF3B6/2020\|EPI_ISL_559839\|20 | ···················· | ···················· | ···················· | ···················· | ···················· | ···················· |
| J | 1519 hCoV-19/England/QEUH-9AF44A/2020\|EPI_ISL_559841\|20 | ···················· | ···················· | ···················· | ···················· | ···················· | ···················· |
| J | 1520 hCoV-19/England/QEUH-9AF459/2020\|EPI_ISL_559842\|20 | ···················· | ···················· | ···················· | ···················· | ···················· | ···················· |
| J | 1521 hCoV-19/England/QEUH-9AF486/2020\|EPI_ISL_559843\|20 | ···················· | ···················· | ···················· | ···················· | ···················· | ···················· |
| J | 1522 hCoV-19/England/QEUH-9AF495/2020\|EPI_ISL_559844\|20 | ···················· | ···················· | ···················· | ···················· | ···················· | ···················· |
| J | 1523 hCoV-19/England/QEUH-9AF4A4/2020\|EPI_ISL_559845\|20 | ···················· | ···················· | ···················· | ···················· | ···················· | ···················· |
| J | 1524 hCoV-19/England/QEUH-9AF4C2/2020\|EPI_ISL_559847\|20 | ···················· | ···················· | ···················· | ···················· | ···················· | ···················· |
| J | 1525 hCoV-19/England/QEUH-9AF4D1/2020\|EPI_ISL_559848\|20 | ···················· | ···················· | ···················· | ···················· | ···················· | ···················· |
| J | 1526 hCoV-19/England/QEUH-9AF529/2020\|EPI_ISL_559849\|20 | ···················· | ···················· | ···················· | ···················· | ···················· | ···················· |
| J | 1527 hCoV-19/England/QEUH-9AF592/2020\|EPI_ISL_559850\|20 | ···················· | ···················· | ···················· | ···················· | ···················· | ···················· |
| J | 1528 hCoV-19/England/QEUH-9AF5DE/2020\|EPI_ISL_559851\|20 | ···················· | ···················· | ···················· | ···················· | ···················· | ···················· |
| J | 1529 hCoV-19/England/QEUH-9AF608/2020\|EPI_ISL_559852\|20 | ···················· | ···················· | ···················· | ···················· | ···················· | ···················· |
| J | 1530 hCoV-19/England/QEUH-9AF617/2020\|EPI_ISL_559853\|20 | ···················· | ···················· | ···················· | ···················· | ···················· | ···················· |
| J | 1531 hCoV-19/England/QEUH-9AF644/2020\|EPI_ISL_559854\|20 | ···················· | ···················· | ···················· | ···················· | ···················· | ···················· |
| J | 1532 hCoV-19/England/QEUH-9B0BAB/2020\|EPI_ISL_559855\|20 | ···················· | ···················· | ···················· | ···················· | ···················· | ···················· |
| J | 1533 hCoV-19/England/QEUH-9B0C5D/2020\|EPI_ISL_559856\|20 | ···················· | ···················· | ···················· | ···················· | ···················· | ···················· |
| J | 1534 hCoV-19/England/QEUH-9B0C6C/2020\|EPI_ISL_559857\|20 | ···················· | ···················· | ···················· | ···················· | ······T············· | ···················· |
| J | 1535 hCoV-19/England/QEUH-9B42DF/2020\|EPI_ISL_559888\|20 | ···················· | ···················· | ···················· | ···················· | ···················· | ···················· |
| J | 1536 hCoV-19/England/QEUH-9B43AF/2020\|EPI_ISL_559889\|20 | ···················· | ···················· | ···················· | ···················· | ···················· | ···················· |
| J | 1537 hCoV-19/England/MILK-9AAFE4/2020\|EPI_ISL_551124\|20 | ···················· | ·A·················· | ···················· | ···················· | ···················· | ···A················ |
| J | 1538 hCoV-19/England/MILK-9A9598/2020\|EPI_ISL_551126\|20 | ···················· | ···················· | ···················· | ···················· | ···················· | ···················· |
| J | 1539 hCoV-19/England/MILK-9A945F/2020\|EPI_ISL_551127\|20 | ···················· | ···················· | ···················· | ···················· | ······T············· | ···················· |
| J | 1540 hCoV-19/England/MILK-9A9ED9/2020\|EPI_ISL_551128\|20 | ···················· | ···················· | ···················· | ···················· | ···················· | ···················· |
| J | 1541 hCoV-19/England/MILK-9A9000/2020\|EPI_ISL_551129\|20 | ···················· | ···················· | ···················· | ···················· | ···················· | ···················· |
| J | 1542 hCoV-19/England/MILK-9AB499/2020\|EPI_ISL_551130\|20 | ···················· | ···················· | ···················· | ···················· | ···················· | ···················· |
| J | 1543 hCoV-19/England/MILK-9A9C0C/2020\|EPI_ISL_551135\|20 | ···················· | ···················· | ···················· | ···················· | ···················· | ···················· |
| J | 1544 hCoV-19/England/MILK-9A8E34/2020\|EPI_ISL_551136\|20 | ···················· | ···················· | ···················· | ···················· | ···················· | ···················· |
| J | 1545 hCoV-19/England/MILK-9A9B96/2020\|EPI_ISL_551138\|20 | ···················· | ···················· | ···················· | ···················· | ···················· | ···················· |
| J | 1546 hCoV-19/England/MILK-9A9C75/2020\|EPI_ISL_551139\|20 | ···················· | ···················· | ···················· | ···················· | ···················· | ···················· |
| J | 1547 hCoV-19/England/MILK-9A9440/2020\|EPI_ISL_551140\|20 | ···················· | ·A·················· | ···················· | ···················· | ···················· | ···················· |
| J | 1548 hCoV-19/England/MILK-9A9361/2020\|EPI_ISL_551141\|20 | ···················· | ···················· | ···················· | ···················· | ···················· | ···················· |
| J | 1549 hCoV-19/England/MILK-9AB10B/2020\|EPI_ISL_551142\|20 | ···················· | ·A·················· | ···················· | ···················· | ···················· | ···A················ |
| J | 1550 hCoV-19/England/MILK-9A961D/2020\|EPI_ISL_551143\|20 | ···················· | ···················· | ···················· | ···················· | ···················· | ···················· |
| J | 1551 hCoV-19/England/MILK-9A905B/2020\|EPI_ISL_551144\|20 | ···················· | ···················· | ···················· | ···················· | ···················· | ···················· |
| J | 1552 hCoV-19/England/MILK-9A9273/2020\|EPI_ISL_551145\|20 | ···················· | ···················· | ···················· | ···················· | ···················· | ···················· |
| J | 1553 hCoV-19/England/MILK-9A906A/2020\|EPI_ISL_551146\|20 | ···················· | ···················· | ···················· | ···················· | ···················· | ···················· |
| J | 1554 hCoV-19/England/MILK-9A8F31/2020\|EPI_ISL_551147\|20 | ···················· | ···················· | ···················· | ···················· | ···················· | ···················· |
| J | 1555 hCoV-19/England/MILK-9A8FB9/2020\|EPI_ISL_551148\|20 | ···················· | ···················· | ···················· | ···················· | ···················· | ···················· |
| J | 1556 hCoV-19/England/MILK-9A903D/2020\|EPI_ISL_551149\|20 | ···················· | ···················· | ···················· | ···················· | ···················· | ···················· |
| J | 1557 hCoV-19/England/MILK-9A912B/2020\|EPI_ISL_551150\|20 | ···················· | ···················· | ···················· | ···················· | ···················· | ···················· |
| J | 1558 hCoV-19/England/MILK-9AB44E/2020\|EPI_ISL_551151\|20 | ···················· | ···················· | ···················· | ···················· | ···················· | ···················· |
| J | 1559 hCoV-19/England/MILK-9A9370/2020\|EPI_ISL_551152\|20 | ···················· | ···················· | ···················· | ···················· | ···················· | ···················· |
| J | 1560 hCoV-19/England/MILK-9A96D1/2020\|EPI_ISL_551153\|20 | ···················· | ···················· | ···················· | ···················· | ···················· | ···················· |
| J | 1561 hCoV-19/England/MILK-9AB165/2020\|EPI_ISL_551154\|20 | ···················· | ·A·················· | ···················· | ···················· | ···················· | ···A················ |
| J | 1562 hCoV-19/England/MILK-9A9194/2020\|EPI_ISL_551155\|20 | ···················· | ···················· | ···················· | ···················· | ···················· | ···················· |
| J | 1563 hCoV-19/England/MILK-9A9E51/2020\|EPI_ISL_551156\|20 | ···················· | ···················· | ···················· | ···················· | ···················· | ···················· |
| J | 1564 hCoV-19/England/MILK-9A9747/2020\|EPI_ISL_551157\|20 | ···················· | ···················· | ···················· | ···················· | ···················· | ···················· |
| J | 1565 hCoV-19/England/MILK-9AAE32/2020\|EPI_ISL_551158\|20 | ···················· | ···················· | ···················· | ···················· | ···················· | ···················· |
| J | 1566 hCoV-19/England/MILK-9A9C84/2020\|EPI_ISL_551161\|20 | ···················· | ···················· | ···················· | ···················· | ···················· | ···················· |
| J | 1567 hCoV-19/England/MILK-9AB4C6/2020\|EPI_ISL_551162\|20 | ···················· | ···················· | ···················· | ···················· | ···················· | ···················· |
| J | 1568 hCoV-19/England/MILK-9A95F2/2020\|EPI_ISL_551163\|20 | ···················· | ···················· | ···················· | ···················· | ······T············· | ···················· |
| J | 1569 hCoV-19/England/MILK-9A90A6/2020\|EPI_ISL_551164\|20 | ···················· | ···················· | ···················· | ···················· | ···················· | ···················· |
| J | 1570 hCoV-19/England/MILK-9A956B/2020\|EPI_ISL_551165\|20 | ···················· | ···················· | ···················· | ···················· | ···················· | ···················· |
| J | 1571 hCoV-19/Wales/MILK-9A9255/2020\|EPI_ISL_551166\|2020 | ···················· | ···················· | ···················· | ···················· | ···················· | ···················· |
| J | 1572 hCoV-19/England/MILK-9A9167/2020\|EPI_ISL_551167\|20 | ···················· | ···················· | ···················· | ···················· | ···················· | ···················· |
| J | 1573 hCoV-19/England/MILK-9A92EC/2020\|EPI_ISL_551168\|20 | ···················· | ···················· | ···················· | ···················· | ···················· | ···················· |
| J | 1574 hCoV-19/England/MILK-9A9C2A/2020\|EPI_ISL_551169\|20 | ···················· | ···················· | ···················· | ···················· | ···················· | ···················· |
| J | 1575 hCoV-19/England/MILK-9A9CFD/2020\|EPI_ISL_551170\|20 | ···················· | ·A·················· | ···················· | ···················· | ···················· | ···A················ |

| | | | | | | | | |
|---|---|---|---|---|---|---|---|---|
| J | 1576 | hCoV-19/England/MILK-9A9079/2020\|EPI_ISL_551171\|20 | ···················· | ···················· | ···················· | ···················· | ···················· | ···················· |
| J | 1577 | hCoV-19/England/MILK-9A9783/2020\|EPI_ISL_551172\|20 | ···················· | ···················· | ···················· | ···················· | ···················· | ···················· |
| J | 1578 | hCoV-19/England/MILK-9AB2BD/2020\|EPI_ISL_551173\|20 | ···················· | ···················· | ···················· | ···················· | ···················· | ···················· |
| J | 1579 | hCoV-19/England/MILK-9A9B0F/2020\|EPI_ISL_551174\|20 | ···················· | ···················· | ···················· | ···················· | ···················· | ···················· |
| J | 1580 | hCoV-19/England/MILK-9A9A20/2020\|EPI_ISL_551175\|20 | ···················· | ·A·················· | ···················· | ···················· | ···················· | ··A················· |
| J | 1581 | hCoV-19/England/MILK-9AAE41/2020\|EPI_ISL_551176\|20 | ···················· | ···················· | ···················· | ···················· | ···················· | ···················· |
| J | 1582 | hCoV-19/England/MILK-9A949B/2020\|EPI_ISL_551178\|20 | ···················· | ···················· | ···················· | ···················· | ···················· | ···················· |
| J | 1583 | hCoV-19/England/MILK-991CCB/2020\|EPI_ISL_552400\|20 | ···················· | ···················· | ···················· | ···················· | ···················· | ···················· |
| J | 1584 | hCoV-19/England/MILK-993968/2020\|EPI_ISL_552401\|20 | ···················· | ···················· | ···················· | ···················· | ···················· | ···················· |
| J | 1585 | hCoV-19/England/MILK-994FA1/2020\|EPI_ISL_552402\|20 | ···················· | ···················· | ···················· | ···················· | ···················· | ···················· |
| J | 1586 | hCoV-19/England/MILK-99515F/2020\|EPI_ISL_552405\|20 | ···················· | ···················· | ···················· | ···················· | ···················· | ···················· |
| J | 1587 | hCoV-19/England/MILK-993ADE/2020\|EPI_ISL_552406\|20 | ···················· | ···················· | ···················· | ···················· | ···················· | ···················· |
| J | 1588 | hCoV-19/England/MILK-99532C/2020\|EPI_ISL_552407\|20 | ···················· | ···················· | ···················· | ···················· | ···················· | ···················· |
| J | 1589 | hCoV-19/Wales/MILK-99530E/2020\|EPI_ISL_552408\|2020 | ···················· | ···················· | ···················· | ···················· | ···················· | ···················· |
| J | 1590 | hCoV-19/England/MILK-991DF5/2020\|EPI_ISL_552409\|20 | ···················· | ···················· | ···················· | ···················· | ···················· | ···················· |
| J | 1591 | hCoV-19/England/MILK-994976/2020\|EPI_ISL_552410\|20 | ···················· | ···················· | ···················· | ···················· | ···················· | ···················· |
| J | 1592 | hCoV-19/England/MILK-9952F2/2020\|EPI_ISL_552411\|20 | ···················· | ···················· | ···················· | ···················· | ···················· | ···················· |
| J | 1593 | hCoV-19/England/MILK-993D99/2020\|EPI_ISL_552412\|20 | ···················· | ···················· | ···················· | ···················· | ···················· | ···················· |
| J | 1594 | hCoV-19/Wales/MILK-994FFC/2020\|EPI_ISL_552414\|2020 | ···················· | ···················· | ···················· | ···················· | ···················· | ···················· |
| J | 1595 | hCoV-19/England/MILK-9953D1/2020\|EPI_ISL_552416\|20 | ···················· | ···················· | ···················· | ···················· | ···················· | ···················· |
| J | 1596 | hCoV-19/Wales/MILK-9950AD/2020\|EPI_ISL_552417\|2020 | ···················· | ···················· | ···················· | ···················· | ···················· | ···················· |
| J | 1597 | hCoV-19/England/MILK-99491C/2020\|EPI_ISL_552418\|20 | ···················· | ···················· | ···················· | ···················· | ···················· | ···················· |
| J | 1598 | hCoV-19/England/MILK-994AFB/2020\|EPI_ISL_552419\|20 | ···················· | ···················· | ···················· | ···················· | ···················· | ···················· |
| J | 1599 | hCoV-19/England/MILK-994FCF/2020\|EPI_ISL_552420\|20 | ···················· | ·A·················· | ···················· | ···················· | ···················· | ··A················· |
| J | 1600 | hCoV-19/England/MILK-9947C7/2020\|EPI_ISL_552421\|20 | ···················· | ···················· | ···················· | ···················· | ···················· | ···················· |
| J | 1601 | hCoV-19/England/MILK-97500D/2020\|EPI_ISL_552455\|20 | ···················· | ···················· | ···················· | ···················· | ···················· | ···················· |
| J | 1602 | hCoV-19/England/MILK-975832/2020\|EPI_ISL_552456\|20 | ···················· | ···················· | ···················· | ···················· | ···················· | ···················· |
| J | 1603 | hCoV-19/England/MILK-9756CF/2020\|EPI_ISL_552459\|20 | ···················· | ·A·················· | ···················· | ···················· | ···················· | ··A················· |
| J | 1604 | hCoV-19/England/MILK-974D43/2020\|EPI_ISL_552460\|20 | ···················· | ···················· | ···················· | ···················· | ···················· | ···················· |
| J | 1605 | hCoV-19/England/MILK-974AB5/2020\|EPI_ISL_552461\|20 | ···················· | ···················· | ···················· | ···················· | ···················· | ···················· |
| J | 1606 | hCoV-19/England/ALDP-98293F/2020\|EPI_ISL_552466\|20 | ···················· | ·A·················· | ···················· | ···················· | ···················· | ···················· |
| J | 1607 | hCoV-19/England/MILK-97545C/2020\|EPI_ISL_552467\|20 | ···················· | ···················· | ···················· | ···················· | ···················· | ···················· |
| J | 1608 | hCoV-19/England/ALDP-982A78/2020\|EPI_ISL_552468\|20 | ···················· | ·A·················· | ···················· | ···················· | ···················· | ··A················· |
| J | 1609 | hCoV-19/England/MILK-975A5A/2020\|EPI_ISL_552469\|20 | ···················· | ···················· | ···················· | ···················· | ···················· | ···················· |
| J | 1610 | hCoV-19/England/MILK-97553B/2020\|EPI_ISL_552470\|20 | ···················· | ···················· | ···················· | ···················· | ···················· | ···················· |
| J | 1611 | hCoV-19/England/MILK-974D34/2020\|EPI_ISL_552471\|20 | ···················· | ···················· | ···················· | ···················· | ···················· | ···················· |
| J | 1612 | hCoV-19/England/MILK-975C18/2020\|EPI_ISL_552472\|20 | ···················· | ···················· | ···················· | ···················· | ···················· | ···················· |
| J | 1613 | hCoV-19/England/ALDP-982D7F/2020\|EPI_ISL_552473\|20 | ···················· | ···················· | ···················· | ···················· | ···················· | ···················· |
| J | 1614 | hCoV-19/England/MILK-97560B/2020\|EPI_ISL_552474\|20 | ···················· | ···················· | ···················· | ···················· | ···················· | ···················· |
| J | 1615 | hCoV-19/England/MILK-975A1E/2020\|EPI_ISL_552475\|20 | ···················· | ···················· | ···················· | ···················· | ···················· | ···················· |
| J | 1616 | hCoV-19/England/ALDP-982BB1/2020\|EPI_ISL_552476\|20 | ···················· | ···················· | ···················· | ···················· | ···················· | ···················· |
| J | 1617 | hCoV-19/England/MILK-975A0F/2020\|EPI_ISL_552477\|20 | ···················· | ···················· | ···················· | ···················· | ···················· | ···················· |
| J | 1618 | hCoV-19/England/MILK-974F3E/2020\|EPI_ISL_552478\|20 | ···················· | ···················· | ···················· | ···················· | ···················· | ···················· |
| J | 1619 | hCoV-19/England/ALDP-982BC0/2020\|EPI_ISL_552479\|20 | ···················· | ···················· | ···················· | ···················· | ···················· | ···················· |
| J | 1620 | hCoV-19/England/MILK-9752E9/2020\|EPI_ISL_552480\|20 | ···················· | ···················· | ···················· | ···················· | ···················· | ···················· |
| J | 1621 | hCoV-19/England/ALDP-9818D9/2020\|EPI_ISL_552481\|20 | ···················· | ···················· | ···················· | ···················· | ···················· | ···················· |
| J | 1622 | hCoV-19/England/MILK-9758AB/2020\|EPI_ISL_552483\|20 | ···················· | ···················· | ···················· | ···················· | ···················· | ···················· |
| J | 1623 | hCoV-19/England/MILK-974D9E/2020\|EPI_ISL_552484\|20 | ···················· | ···················· | ···················· | ···················· | ···················· | ···················· |
| J | 1624 | hCoV-19/England/ALDP-982DAC/2020\|EPI_ISL_552486\|20 | ···················· | ···················· | ···················· | ···················· | ···················· | ···················· |
| J | 1625 | hCoV-19/England/MILK-974DAD/2020\|EPI_ISL_552487\|20 | ···················· | ···················· | ···················· | ···················· | ···················· | ···················· |
| J | 1626 | hCoV-19/England/ALDP-98286F/2020\|EPI_ISL_552488\|20 | ···················· | ···················· | ···················· | ···················· | ···················· | ···················· |
| J | 1627 | hCoV-19/England/MILK-975A87/2020\|EPI_ISL_552490\|20 | ···················· | ···················· | ···················· | ···················· | ···················· | ···················· |
| J | 1628 | hCoV-19/England/MILK-97594E/2020\|EPI_ISL_552491\|20 | ···················· | ···················· | ···················· | ···················· | ···················· | ···················· |
| J | 1629 | hCoV-19/England/MILK-974FA7/2020\|EPI_ISL_552493\|20 | ···················· | ···················· | ···················· | ···················· | ···················· | ···················· |
| J | 1630 | hCoV-19/England/MILK-974ED7/2020\|EPI_ISL_552494\|20 | ···················· | ···················· | ···················· | ···················· | ···················· | ···················· |
| J | 1631 | hCoV-19/England/MILK-97593F/2020\|EPI_ISL_552495\|20 | ···················· | ···················· | ···················· | ···················· | ···················· | ···················· |
| J | 1632 | hCoV-19/Wales/MILK-974B2B/2020\|EPI_ISL_552496\|2020 | ···················· | ···················· | ···················· | ···················· | ···················· | ···················· |
| J | 1633 | hCoV-19/England/MILK-97561A/2020\|EPI_ISL_552497\|20 | ···················· | ···················· | ···················· | ···················· | ···················· | ···················· |
| J | 1634 | hCoV-19/England/MILK-97539B/2020\|EPI_ISL_552498\|20 | ···················· | ···················· | ···················· | ···················· | ···················· | ···················· |
| J | 1635 | hCoV-19/England/MILK-975216/2020\|EPI_ISL_552499\|20 | ···················· | ···················· | ···················· | ···················· | ···················· | ···················· |
| J | 1636 | hCoV-19/England/ALDP-905E26/2020\|EPI_ISL_553702\|20 | ···················· | ···················· | ···················· | ···················· | ···················· | ···················· |
| J | 1637 | hCoV-19/England/ALDP-91BACF/2020\|EPI_ISL_553705\|20 | ···················· | ···················· | ···················· | ···················· | ···················· | ···················· |
| J | 1638 | hCoV-19/England/ALDP-93A875/2020\|EPI_ISL_553707\|20 | ···················· | ···················· | ···················· | ···················· | ···················· | ···················· |
| J | 1639 | hCoV-19/England/ALDP-919F4A/2020\|EPI_ISL_553708\|20 | ···················· | ···················· | ···················· | ···················· | ···················· | ···················· |
| J | 1640 | hCoV-19/England/ALDP-92C102/2020\|EPI_ISL_553709\|20 | ···················· | ···················· | ···················· | ···················· | ···················· | ···················· |
| J | 1641 | hCoV-19/England/ALDP-91BD11/2020\|EPI_ISL_553710\|20 | ···················· | ···················· | ···················· | ···················· | ···················· | ···················· |
| J | 1642 | hCoV-19/England/ALDP-92AE3A/2020\|EPI_ISL_553711\|20 | ···················· | ···················· | ···················· | ···················· | ···················· | ···················· |
| J | 1643 | hCoV-19/Wales/ALDP-90E2A9/2020\|EPI_ISL_553712\|2020 | ···················· | ···················· | ···················· | ···················· | ···················· | ···················· |
| J | 1644 | hCoV-19/England/ALDP-9047B3/2020\|EPI_ISL_553713\|20 | ···················· | ···················· | ···················· | ···················· | ···················· | ···················· |
| J | 1645 | hCoV-19/England/ALDP-924E12/2020\|EPI_ISL_553717\|20 | ···················· | ···················· | ···················· | ···················· | ···················· | ···················· |
| J | 1646 | hCoV-19/England/ALDP-90D389/2020\|EPI_ISL_553718\|20 | ···················· | ···················· | ···················· | ···················· | ···················· | ···················· |
| J | 1647 | hCoV-19/England/ALDP-916DDA/2020\|EPI_ISL_553720\|20 | ···················· | ···················· | ···················· | ···················· | ···················· | ···················· |
| J | 1648 | hCoV-19/England/ALDP-924D60/2020\|EPI_ISL_553721\|20 | ···················· | ···················· | ···················· | ···················· | ···················· | ···················· |
| J | 1649 | hCoV-19/England/ALDP-91073C/2020\|EPI_ISL_553723\|20 | ···················· | ···················· | ···················· | ···················· | ···················· | ···················· |
| J | 1650 | hCoV-19/England/ALDP-92ACB8/2020\|EPI_ISL_553724\|20 | ···················· | ···················· | ···················· | ···················· | ···················· | ···················· |
| J | 1651 | hCoV-19/England/ALDP-9067DF/2020\|EPI_ISL_553725\|20 | ···················· | ···················· | ···················· | ···················· | ···················· | ···················· |
| J | 1652 | hCoV-19/England/ALDP-909F3E/2020\|EPI_ISL_553729\|20 | ···················· | ···················· | ···················· | ···················· | ···················· | ···················· |
| J | 1653 | hCoV-19/England/ALDP-90C86D/2020\|EPI_ISL_553730\|20 | ···················· | ·A·················· | ···················· | ···················· | ···················· | ···················· |
| J | 1654 | hCoV-19/England/QEUH-96AFFF/2020\|EPI_ISL_553732\|20 | ···················· | ···················· | ···················· | ···················· | ···················· | ···················· |
| J | 1655 | hCoV-19/England/ALDP-9543FC/2020\|EPI_ISL_553733\|20 | ···················· | ···················· | ···················· | ···················· | ···················· | ···················· |

J 1656 hCoV-19/England/ALDP-954189/2020|EPI_ISL_553735|20 ···················· ···················· ···················· ···················· ···················· ····················
J 1657 hCoV-19/England/ALDP-954110/2020|EPI_ISL_553736|20 ···················· ···················· ···················· ···················· ···················· ····················
J 1658 hCoV-19/England/ALDP-9541D4/2020|EPI_ISL_553737|20 ···················· ···················· ···················· ···················· ···················· ····················
J 1659 hCoV-19/England/ALDP-953A07/2020|EPI_ISL_553739|20 ···················· ···················· ···················· ···················· ···················· ····················
J 1660 hCoV-19/England/ALDP-95459C/2020|EPI_ISL_553740|20 ···················· ···················· ···················· ···················· ···················· ····················
J 1661 hCoV-19/England/ALDP-9537C4/2020|EPI_ISL_553742|20 ···················· ···················· ···················· ···················· ···················· ····················
J 1662 hCoV-19/England/ALDP-9539EC/2020|EPI_ISL_553744|20 ···················· ···················· ···················· ···················· ···················· ····················
J 1663 hCoV-19/England/ALDP-954338/2020|EPI_ISL_553745|20 ···················· ···················· ···················· ···················· ···················· ····················
J 1664 hCoV-19/England/ALDP-954480/2020|EPI_ISL_553747|20 ···················· ···················· ···················· ···················· ···················· ····················
J 1665 hCoV-19/England/ALDP-953B13/2020|EPI_ISL_553748|20 ···················· ···················· ···················· ···················· ···················· ····················
J 1666 hCoV-19/England/ALDP-953A43/2020|EPI_ISL_553749|20 ···················· ···················· ···················· ···················· ···················· ····················
J 1667 hCoV-19/England/ALDP-953BC8/2020|EPI_ISL_553750|20 ···················· ···················· ···················· ···················· ···················· ····················
J 1668 hCoV-19/England/ALDP-953849/2020|EPI_ISL_553751|20 ···················· ···················· ···················· ···················· ···················· ····················
J 1669 hCoV-19/England/ALDP-953603/2020|EPI_ISL_553752|20 ···················· ···················· ···················· ···················· ···················· ····················
J 1670 hCoV-19/England/ALDP-953B40/2020|EPI_ISL_553786|20 ···················· ···················· ···················· ···················· ···················· ····················
J 1671 hCoV-19/England/ALDP-954602/2020|EPI_ISL_553787|20 ···················· ···················· ···················· ···················· ···················· ····················
J 1672 hCoV-19/England/ALDP-954462/2020|EPI_ISL_553788|20 ···················· ···················· ···················· ···················· ···················· ····················
J 1673 hCoV-19/England/ALDP-953560/2020|EPI_ISL_553789|20 ···················· ···················· ···················· ···················· ···················· ····················
J 1674 hCoV-19/England/ALDP-953B04/2020|EPI_ISL_553792|20 ···················· ···················· ···················· ···················· ···················· ····················
J 1675 hCoV-19/England/ALDP-953AF8/2020|EPI_ISL_553797|20 ···················· ···················· ···················· ···················· ···················· ····················
J 1676 hCoV-19/England/ALDP-95422C/2020|EPI_ISL_553798|20 ···················· ···················· ···················· ···················· ···················· ····················
J 1677 hCoV-19/England/ALDP-953E56/2020|EPI_ISL_553799|20 ···················· ·A·················· ···················· ···················· ···················· ····················
J 1678 hCoV-19/England/ALDP-8CF09C/2020|EPI_ISL_557100|20 ···················· ···················· ···················· ···················· ···················· ····················
J 1679 hCoV-19/England/ALDP-8BA43E/2020|EPI_ISL_557101|20 ···················· ···················· ···················· ···················· ···················· ····················
J 1680 hCoV-19/England/ALDP-8FFEF3/2020|EPI_ISL_557102|20 ···················· ···················· ···················· ···················· ···················· ····················
J 1681 hCoV-19/England/ALDP-8FF9A7/2020|EPI_ISL_557104|20 ···················· ···················· ···················· ···················· ···················· ····················
J 1682 hCoV-19/England/ALDP-8D64D2/2020|EPI_ISL_557105|20 ···················· ···················· ···················· ···················· ···················· ····················
J 1683 hCoV-19/England/ALDP-8DEC69/2020|EPI_ISL_557106|20 ···················· ···················· ···················· ···················· ···················· ····················
J 1684 hCoV-19/England/ALDP-8FE067/2020|EPI_ISL_557107|20 ···················· ···················· ···················· ···················· ···················· ····················
J 1685 hCoV-19/England/ALDP-8D741D/2020|EPI_ISL_557108|20 ···················· ···················· ···················· ···················· ···················· ····················
J 1686 hCoV-19/England/ALDP-8E2FF0/2020|EPI_ISL_557109|20 ···················· ···················· ···················· ···················· ···················· ····················
J 1687 hCoV-19/England/ALDP-8B8CFC/2020|EPI_ISL_557111|20 ···················· ···················· ···················· ···················· ···················· ····················
J 1688 hCoV-19/England/ALDP-8B7273/2020|EPI_ISL_557113|20 ···················· ···················· ···················· ···················· ···················· ····················
J 1689 hCoV-19/England/ALDP-8D728C/2020|EPI_ISL_557114|20 ···················· ···················· ···················· ···················· ···················· ····················
J 1690 hCoV-19/England/ALDP-8D3CA0/2020|EPI_ISL_557115|20 ···················· ···················· ···················· ···················· ···················· ····················
J 1691 hCoV-19/England/ALDP-8FE1DD/2020|EPI_ISL_557116|20 ···················· ···················· ···················· ···················· ···················· ····················
J 1692 hCoV-19/England/ALDP-8FDEF5/2020|EPI_ISL_557118|20 ···················· ···················· ···················· ···················· ···················· ····················
J 1693 hCoV-19/England/ALDP-8D40C1/2020|EPI_ISL_557120|20 ···················· ···················· ···················· ···················· ···················· ····················
J 1694 hCoV-19/England/ALDP-8B48B0/2020|EPI_ISL_557121|20 ···················· ···················· ···················· ···················· ···················· ····················
J 1695 hCoV-19/England/ALDP-8BA0A3/2020|EPI_ISL_557122|20 ···················· ···················· ···················· ···················· ···················· ····················
J 1696 hCoV-19/England/ALDP-8E2C26/2020|EPI_ISL_557123|20 ···················· ···················· ···················· ···················· ···················· ····················
J 1697 hCoV-19/England/ALDP-8DD1F2/2020|EPI_ISL_557124|20 ···················· ···················· ···················· ···················· ···················· ····················
J 1698 hCoV-19/England/ALDP-8E2C17/2020|EPI_ISL_557125|20 ···················· ···················· ···················· ···················· ···················· ····················
J 1699 hCoV-19/Wales/ALDP-8D984C/2020|EPI_ISL_557126|2020 ···················· ···················· ···················· ···················· ···················· ····················
J 1700 hCoV-19/England/ALDP-8D4D06/2020|EPI_ISL_557127|20 ···················· ···················· ···················· ···················· ···················· ····················
J 1701 hCoV-19/England/ALDP-8DCF17/2020|EPI_ISL_557128|20 ···················· ···················· ···················· ···················· ···················· ····················
J 1702 hCoV-19/England/MILK-82A8E2/2020|EPI_ISL_557280|20 ···················· ···················· ···················· ···················· ···················· ····················
J 1703 hCoV-19/England/MILK-81D8E2/2020|EPI_ISL_557281|20 ···················· ···················· ···················· ···················· ···················· ····················
J 1704 hCoV-19/England/MILK-800BDA/2020|EPI_ISL_557282|20 ···················· ···················· ···················· ···················· ···················· ····················
J 1705 hCoV-19/England/MILK-848286/2020|EPI_ISL_557285|20 ···················· ·A·················· ···················· ···················· ···················· ····················
J 1706 hCoV-19/England/MILK-8781DB/2020|EPI_ISL_557286|20 ···················· ···················· ···················· ···················· ···················· ····················
J 1707 hCoV-19/England/MILK-82154E/2020|EPI_ISL_557288|20 ···················· ···················· ···················· ···················· ···················· ····················
J 1708 hCoV-19/England/MILK-847DFF/2020|EPI_ISL_557289|20 ···················· ···················· ···················· ···················· ···················· ····················
J 1709 hCoV-19/England/MILK-858247/2020|EPI_ISL_557290|20 ···················· ···················· ···················· ···················· ···················· ····················
J 1710 hCoV-19/England/MILK-8313C9/2020|EPI_ISL_557291|20 ···················· ···················· ···················· ···················· ···················· ····················
J 1711 hCoV-19/England/MILK-846947/2020|EPI_ISL_557292|20 ···················· ···················· ···················· ···················· ···················· ····················
J 1712 hCoV-19/England/MILK-830E9C/2020|EPI_ISL_557293|20 ···················· ···················· ···················· ···················· ···················· ····················
J 1713 hCoV-19/England/MILK-8477D3/2020|EPI_ISL_557296|20 ···················· ···················· ···················· ···················· ···················· ····················
J 1714 hCoV-19/England/MILK-84490D/2020|EPI_ISL_557297|20 ···················· ···················· ···················· ···················· ···················· ····················
J 1715 hCoV-19/England/MILK-818DBB/2020|EPI_ISL_557298|20 ···················· ···················· ···················· ···················· ···················· ····················
J 1716 hCoV-19/England/ALDP-5D04BD/2020|EPI_ISL_558406|20 ···················· ···················· ···················· ···················· ···················· ····················
J 1717 hCoV-19/England/ALDP-5C7E9D/2020|EPI_ISL_558407|20 ···················· ···················· ···················· ···················· ···················· ····················
J 1718 hCoV-19/England/ALDP-5CAAB4/2020|EPI_ISL_558409|20 ···················· ···················· ···················· ···················· ···················· ····················
J 1719 hCoV-19/England/ALDP-5C70D3/2020|EPI_ISL_558410|20 ···················· ···················· ···················· ···················· ···················· ····················
J 1720 hCoV-19/England/ALDP-5D1B02/2020|EPI_ISL_558411|20 ···················· ·A·················· ···················· ···················· ···················· ····················
J 1721 hCoV-19/England/ALDP-5D44B9/2020|EPI_ISL_558412|20 ···················· ···················· ···················· ···················· ···················· ····················
J 1722 hCoV-19/England/ALDP-5D3818/2020|EPI_ISL_558413|20 ···················· ···················· ···················· ···················· ···················· ····················
J 1723 hCoV-19/England/ALDP-5C9068/2020|EPI_ISL_558414|20 ···················· ···················· ···················· ···················· ···················· ····················
J 1724 hCoV-19/England/ALDP-5D307A/2020|EPI_ISL_558416|20 ···················· ···················· ···················· ···················· ···················· ····················
J 1725 hCoV-19/England/ALDP-5D65C3/2020|EPI_ISL_558417|20 ···················· ···················· ···················· ···················· ···················· ····················
J 1726 hCoV-19/England/ALDP-5C998B/2020|EPI_ISL_558418|20 ···················· ···················· ···················· ···················· ···················· ····················
J 1727 hCoV-19/England/ALDP-5D0514/2020|EPI_ISL_558420|20 ···················· ···················· ···················· ···················· ···················· ····················
J 1728 hCoV-19/England/ALDP-5D1829/2020|EPI_ISL_558421|20 ···················· ···················· ···················· ···················· ···················· ····················
J 1729 hCoV-19/England/ALDP-5C9569/2020|EPI_ISL_558423|20 ···················· ···················· ···················· ···················· ···················· ····················
J 1730 hCoV-19/England/ALDP-5C705B/2020|EPI_ISL_558424|20 ···················· ···················· ···················· ···················· ···················· ····················
J 1731 hCoV-19/England/ALDP-5D4C84/2020|EPI_ISL_558425|20 ···················· ···················· ···················· ···················· ···················· ····················
J 1732 hCoV-19/England/ALDP-5D4B1E/2020|EPI_ISL_558426|20 ···················· ···················· ···················· ···················· ···················· ····················
J 1733 hCoV-19/England/ALDP-5CB136/2020|EPI_ISL_558427|20 ···················· ···················· ···················· ···················· ···················· ····················
J 1734 hCoV-19/England/ALDP-5C6D55/2020|EPI_ISL_558429|20 ···················· ···················· ···················· ···················· ···················· ····················
J 1735 hCoV-19/England/ALDP-5D9A1C/2020|EPI_ISL_558430|20 ···················· ···················· ···················· ···················· ···················· ····················

| | | | | | | | | |
|---|---|---|---|---|---|---|---|---|
| J | 1736 | hCoV-19/England/ALDP-5D5F5D/2020\|EPI_ISL_558431\|20 | ···················· | ···················· | ···················· | ···················· | ···················· | ···················· |
| J | 1737 | hCoV-19/England/ALDP-5D05D8/2020\|EPI_ISL_558433\|20 | ···················· | ···················· | ···················· | ···················· | ···················· | ···················· |
| J | 1738 | hCoV-19/England/ALDP-5D3696/2020\|EPI_ISL_558435\|20 | ···················· | ···················· | ···················· | ···················· | ···················· | ···················· |
| J | 1739 | hCoV-19/England/ALDP-5C8834/2020\|EPI_ISL_558436\|20 | ···················· | ···················· | ···················· | ···················· | ···················· | ···················· |
| J | 1740 | hCoV-19/England/ALDP-5C756B/2020\|EPI_ISL_558438\|20 | ···················· | ···················· | ···················· | ···················· | ···················· | ···················· |
| J | 1741 | hCoV-19/England/ALDP-5C51E1/2020\|EPI_ISL_558439\|20 | ···················· | ···················· | ···················· | ···················· | ···················· | ···················· |
| J | 1742 | hCoV-19/England/ALDP-5CC9A6/2020\|EPI_ISL_558442\|20 | ···················· | ···················· | ···················· | ···················· | ···················· | ···················· |
| J | 1743 | hCoV-19/England/ALDP-5CEC32/2020\|EPI_ISL_558443\|20 | ···················· | ···················· | ···················· | ···················· | ···················· | ···················· |
| J | 1744 | hCoV-19/England/ALDP-5C7EAC/2020\|EPI_ISL_558445\|20 | ···················· | ···················· | ···················· | ···················· | ···················· | ···················· |
| J | 1745 | hCoV-19/England/ALDP-5CA7F9/2020\|EPI_ISL_558447\|20 | ···················· | ···················· | ···················· | ···················· | ···················· | ···················· |
| J | 1746 | hCoV-19/England/ALDP-5C9A88/2020\|EPI_ISL_558448\|20 | ···················· | ···················· | ···················· | ···················· | ···················· | ···················· |
| J | 1747 | hCoV-19/England/ALDP-5D0A42/2020\|EPI_ISL_558449\|20 | ···················· | ···················· | ···················· | ···················· | ···················· | ···················· |
| J | 1748 | hCoV-19/England/ALDP-5CCD22/2020\|EPI_ISL_558460\|20 | ···················· | ···················· | ···················· | ···················· | ···················· | ···················· |
| J | 1749 | hCoV-19/England/ALDP-5CF475/2020\|EPI_ISL_558461\|20 | ···················· | ···················· | ···················· | ···················· | ···················· | ···················· |
| J | 1750 | hCoV-19/England/MILK-467B7B/2020\|EPI_ISL_559737\|20 | ···················· | ···················· | ···················· | ···················· | ···················· | ···················· |
| J | 1751 | hCoV-19/England/MILK-4691FF/2020\|EPI_ISL_559738\|20 | ···················· | ···················· | ···················· | ···················· | ···················· | ···················· |
| J | 1752 | hCoV-19/England/MILK-467610/2020\|EPI_ISL_559739\|20 | ···················· | ···················· | ···················· | ···················· | ···················· | ···················· |
| J | 1753 | hCoV-19/England/MILK-460969/2020\|EPI_ISL_559741\|20 | ···················· | ···················· | ···················· | ···················· | ···················· | ···················· |
| J | 1754 | hCoV-19/England/MILK-460D5E/2020\|EPI_ISL_559742\|20 | ···················· | ···················· | ···················· | ···················· | ···················· | ···················· |
| J | 1755 | hCoV-19/Wales/MILK-45F7FA/2020\|EPI_ISL_559743\|2020 | ···················· | ···················· | ···················· | ···················· | ···················· | ···················· |
| J | 1756 | hCoV-19/England/MILK-45F675/2020\|EPI_ISL_559744\|20 | ···················· | ···················· | ···················· | ···················· | ···················· | ···················· |
| J | 1757 | hCoV-19/England/MILK-45EAD4/2020\|EPI_ISL_559745\|20 | ···················· | ···················· | ···················· | ···················· | ···················· | ···················· |
| J | 1758 | hCoV-19/England/MILK-4694C9/2020\|EPI_ISL_559746\|20 | ···················· | ···················· | ···················· | ···················· | ···················· | ···················· |
| J | 1759 | hCoV-19/England/MILK-46958A/2020\|EPI_ISL_559747\|20 | ···················· | ···················· | ···················· | ···················· | ···················· | ···················· |
| J | 1760 | hCoV-19/England/MILK-466A15/2020\|EPI_ISL_559749\|20 | ···················· | ···················· | ···················· | ···················· | ···················· | ···················· |
| J | 1761 | hCoV-19/England/MILK-46667B/2020\|EPI_ISL_559750\|20 | ···················· | ···················· | ···················· | ···················· | ···················· | ···················· |
| J | 1762 | hCoV-19/England/MILK-460C7F/2020\|EPI_ISL_559751\|20 | ···················· | ···················· | ···················· | ···················· | ···················· | ···················· |
| J | 1763 | hCoV-19/England/QEUH-997C2F/2020\|EPI_ISL_559753\|20 | ···················· | ···················· | ···················· | ···················· | ····T··············· | ···················· |
| J | 1764 | hCoV-19/England/QEUH-997C4D/2020\|EPI_ISL_559754\|20 | ···················· | ·A·················· | ···················· | ···················· | ···················· | ···················· |
| J | 1765 | hCoV-19/England/QEUH-997C5C/2020\|EPI_ISL_559755\|20 | ···················· | ···················· | ···················· | ···················· | ····T··············· | ···················· |
| J | 1766 | hCoV-19/England/QEUH-997C7A/2020\|EPI_ISL_559756\|20 | ···················· | ···················· | ···················· | ···················· | ···················· | ···················· |
| J | 1767 | hCoV-19/England/QEUH-997C89/2020\|EPI_ISL_559757\|20 | ···················· | ·A·················· | ···················· | ···················· | ···················· | ···················· |
| J | 1768 | hCoV-19/England/QEUH-997C98/2020\|EPI_ISL_559758\|20 | ···················· | ·A·················· | ···················· | ···················· | ···················· | ···················· |
| J | 1769 | hCoV-19/England/QEUH-997CB6/2020\|EPI_ISL_559759\|20 | ···················· | ···················· | ···················· | ···················· | ······T············· | ···················· |
| J | 1770 | hCoV-19/England/QEUH-997CC5/2020\|EPI_ISL_559760\|20 | ···················· | ···················· | ···················· | ···················· | ···················· | ···················· |
| J | 1771 | hCoV-19/England/QEUH-999470/2020\|EPI_ISL_559761\|20 | ···················· | ···················· | ···················· | ···················· | ···················· | ···················· |
| J | 1772 | hCoV-19/England/QEUH-99948F/2020\|EPI_ISL_559762\|20 | ···················· | ·········A·········· | ···················· | ···················· | ······T············· | ···················· |
| J | 1773 | hCoV-19/England/QEUH-9994AD/2020\|EPI_ISL_559763\|20 | ···················· | ···················· | ···················· | ···················· | ···················· | ···················· |
| J | 1774 | hCoV-19/England/QEUH-9994CB/2020\|EPI_ISL_559764\|20 | ···················· | ·A·················· | ···················· | ···················· | ···················· | ···A················ |
| J | 1775 | hCoV-19/England/QEUH-99B14A/2020\|EPI_ISL_559765\|20 | ···················· | ···················· | ···················· | ···················· | ···················· | ···················· |
| J | 1776 | hCoV-19/England/QEUH-99B159/2020\|EPI_ISL_559766\|20 | ···················· | ···················· | ···················· | ···················· | ····T··············· | ···················· |
| J | 1777 | hCoV-19/England/QEUH-99B168/2020\|EPI_ISL_559767\|20 | ···················· | ···················· | ···················· | ···················· | ····T··············· | ···················· |
| J | 1778 | hCoV-19/England/QEUH-99B8AF/2020\|EPI_ISL_559768\|20 | ···················· | ···················· | ···················· | ···················· | ···················· | ···················· |
| J | 1779 | hCoV-19/England/QEUH-99B933/2020\|EPI_ISL_559769\|20 | ···················· | ·A·················· | ···················· | ···················· | ···················· | ···················· |
| J | 1780 | hCoV-19/England/QEUH-99BC1C/2020\|EPI_ISL_559770\|20 | ···················· | ···················· | ···················· | ···················· | ····T··············· | ···················· |
| J | 1781 | hCoV-19/England/QEUH-99BC3A/2020\|EPI_ISL_559771\|20 | ···················· | ···················· | ···················· | ···················· | ···················· | ···················· |
| J | 1782 | hCoV-19/England/QEUH-99BC49/2020\|EPI_ISL_559772\|20 | ···················· | ···················· | ···················· | ···················· | ···················· | ···················· |
| J | 1783 | hCoV-19/England/QEUH-99BE16/2020\|EPI_ISL_559773\|20 | ···················· | ···················· | ···················· | ···················· | ···················· | ···················· |
| J | 1784 | hCoV-19/England/QEUH-99BE34/2020\|EPI_ISL_559774\|20 | ···················· | ···················· | ···················· | ···················· | ···················· | ···················· |
| J | 1785 | hCoV-19/England/QEUH-99BFAA/2020\|EPI_ISL_559775\|20 | ···················· | ···················· | ···················· | ···················· | ···················· | ···················· |
| J | 1786 | hCoV-19/England/QEUH-99BFB9/2020\|EPI_ISL_559776\|20 | ···················· | ···················· | ···················· | ···················· | ···················· | ···················· |
| J | 1787 | hCoV-19/England/QEUH-99BFD7/2020\|EPI_ISL_559777\|20 | ···················· | ···················· | ···················· | ···················· | ···················· | ···················· |
| J | 1788 | hCoV-19/England/QEUH-99BFE6/2020\|EPI_ISL_559778\|20 | ···················· | ···················· | ···················· | ···················· | ···················· | ···················· |
| J | 1789 | hCoV-19/England/QEUH-99C01F/2020\|EPI_ISL_559779\|20 | ···················· | ···················· | ···················· | ···················· | ····T··············· | ···················· |
| J | 1790 | hCoV-19/England/QEUH-99C282/2020\|EPI_ISL_559790\|20 | ···················· | ···················· | ···················· | ···················· | ···················· | ···················· |
| J | 1791 | hCoV-19/England/QEUH-99C2A0/2020\|EPI_ISL_559791\|20 | ···················· | ···················· | ···················· | ···················· | ···················· | ···············A···· |
| J | 1792 | hCoV-19/England/QEUH-99C2EC/2020\|EPI_ISL_559792\|20 | ···················· | ···················· | ···················· | ···················· | ···················· | ···················· |
| J | 1793 | hCoV-19/England/MILK-9A91EF/2020\|EPI_ISL_551000\|20 | ···················· | ···················· | ···················· | ···················· | ···················· | ···················· |
| J | 1794 | hCoV-19/England/MILK-9A97DE/2020\|EPI_ISL_551002\|20 | ···················· | ···················· | ···················· | ···················· | ···················· | ···················· |
| J | 1795 | hCoV-19/England/MILK-9A8DFB/2020\|EPI_ISL_551004\|20 | ···················· | ···················· | ···················· | ···················· | ···················· | ···················· |
| J | 1796 | hCoV-19/Wales/MILK-9A970B/2020\|EPI_ISL_551005\|2020 | ···················· | ···················· | ···················· | ···················· | ···················· | ···················· |
| J | 1797 | hCoV-19/England/MILK-9A9413/2020\|EPI_ISL_551006\|20 | ···················· | ···················· | ···················· | ···················· | ···················· | ···················· |
| J | 1798 | hCoV-19/England/MILK-9A9291/2020\|EPI_ISL_551008\|20 | ···················· | ···················· | ···················· | ···················· | ···················· | ···················· |
| J | 1799 | hCoV-19/England/MILK-9AB0D1/2020\|EPI_ISL_551009\|20 | ···················· | ···················· | ···················· | ···················· | ···················· | ···················· |
| J | 1800 | hCoV-19/England/MILK-9AB2DB/2020\|EPI_ISL_551010\|20 | ···················· | ···················· | ···················· | ···················· | ···················· | ···················· |
| J | 1801 | hCoV-19/England/MILK-9AAF20/2020\|EPI_ISL_551011\|20 | ···················· | ···················· | ···················· | ···················· | ···················· | ···················· |
| J | 1802 | hCoV-19/England/MILK-9AB37E/2020\|EPI_ISL_551012\|20 | ···················· | ···················· | ···················· | ···················· | ···················· | ···················· |
| J | 1803 | hCoV-19/England/MILK-9A9E42/2020\|EPI_ISL_551014\|20 | ···················· | ···················· | ···················· | ···················· | ···················· | ···················· |
| J | 1804 | hCoV-19/England/MILK-9A9246/2020\|EPI_ISL_551015\|20 | ···················· | ·A·················· | ···················· | ···················· | ···················· | ···A················ |
| J | 1805 | hCoV-19/England/MILK-9A97FC/2020\|EPI_ISL_551016\|20 | ···················· | ···················· | ···················· | ···················· | ···················· | ···················· |
| J | 1806 | hCoV-19/England/MILK-9AB411/2020\|EPI_ISL_551017\|20 | ···················· | ···················· | ···················· | ···················· | ···················· | ···················· |
| J | 1807 | hCoV-19/England/MILK-9AAF3F/2020\|EPI_ISL_551018\|20 | ···················· | ···················· | ···················· | ···················· | ···················· | ···················· |
| J | 1808 | hCoV-19/Wales/MILK-9A913A/2020\|EPI_ISL_551021\|2020 | ···················· | ···················· | ···················· | ···················· | ···················· | ···················· |
| J | 1809 | hCoV-19/England/MILK-9A8EF8/2020\|EPI_ISL_551022\|20 | ···················· | ···················· | ···················· | ···················· | ···················· | ···················· |
| J | 1810 | hCoV-19/England/MILK-9A95D4/2020\|EPI_ISL_551023\|20 | ···················· | ···················· | ···················· | ···················· | ···················· | ···················· |
| J | 1811 | hCoV-19/England/MILK-9A91FE/2020\|EPI_ISL_551024\|20 | ···················· | ···················· | ···················· | ···················· | ·········R·········· | ···················· |
| J | 1812 | hCoV-19/England/MILK-9AAF7B/2020\|EPI_ISL_551033\|20 | ···················· | ···················· | ···················· | ···················· | ···················· | ···················· |
| J | 1813 | hCoV-19/England/MILK-99492B/2020\|EPI_ISL_552268\|20 | ···················· | ···················· | ···················· | ···················· | ···················· | ···················· |
| J | 1814 | hCoV-19/England/MILK-994BE9/2020\|EPI_ISL_552270\|20 | ···················· | ···················· | ···················· | ···················· | ···················· | ···················· |
| J | 1815 | hCoV-19/England/MILK-994A64/2020\|EPI_ISL_552271\|20 | ···················· | ···················· | ···················· | ···················· | ···················· | ···················· |

| | | | | | | | | |
|---|---|---|---|---|---|---|---|---|
| J | 1816 | hCoV-19/England/MILK-993BF9/2020\|EPI_ISL_552272\|20 | ···················· | ···················· | ···················· | ···················· | ···················· | ···················· |
| J | 1817 | hCoV-19/England/MILK-993AED/2020\|EPI_ISL_552273\|20 | ···················· | ···················· | ···················· | ···················· | ···················· | ···················· |
| J | 1818 | hCoV-19/England/MILK-994F29/2020\|EPI_ISL_552275\|20 | ···················· | ···················· | ···················· | ···················· | ···················· | ···················· |
| J | 1819 | hCoV-19/England/MILK-9953A4/2020\|EPI_ISL_552277\|20 | ···················· | ···················· | ···················· | ···················· | ···················· | ···················· |
| J | 1820 | hCoV-19/Wales/MILK-9950DA/2020\|EPI_ISL_552278\|2020 | ···················· | ···················· | ···················· | ···················· | ···················· | ···················· |
| J | 1821 | hCoV-19/England/MILK-993DA8/2020\|EPI_ISL_552279\|20 | ···················· | ···················· | ···················· | ···················· | ···················· | ···················· |
| J | 1822 | hCoV-19/England/MILK-9948F1/2020\|EPI_ISL_552280\|20 | ···················· | ···················· | ···················· | ···················· | ···················· | ···················· |
| J | 1823 | hCoV-19/England/MILK-993CE7/2020\|EPI_ISL_552281\|20 | ···················· | ···················· | ···················· | ···················· | ···················· | ···················· |
| J | 1824 | hCoV-19/England/MILK-99486A/2020\|EPI_ISL_552282\|20 | ···················· | ·A·················· | ···················· | ···················· | ···················· | ···················· |
| J | 1825 | hCoV-19/England/MILK-99518C/2020\|EPI_ISL_552283\|20 | ···················· | ···················· | ···················· | ···················· | ···················· | ···················· |
| J | 1826 | hCoV-19/England/MILK-9939D1/2020\|EPI_ISL_552284\|20 | ···················· | ···················· | ···················· | ···················· | ···················· | ···················· |
| J | 1827 | hCoV-19/England/MILK-9947E5/2020\|EPI_ISL_552285\|20 | ···················· | ···················· | ···················· | ···················· | ···················· | ···················· |
| J | 1828 | hCoV-19/England/MILK-994D01/2020\|EPI_ISL_552287\|20 | ···················· | ···················· | ···················· | ···················· | ···················· | ···················· |
| J | 1829 | hCoV-19/England/MILK-99541A/2020\|EPI_ISL_552288\|20 | ···················· | ···················· | ···················· | ···················· | ···················· | ···················· |
| J | 1830 | hCoV-19/England/MILK-994660/2020\|EPI_ISL_552289\|20 | ···················· | ···················· | ···················· | ···················· | ···················· | ···················· |
| J | 1831 | hCoV-19/England/MILK-991DE6/2020\|EPI_ISL_552290\|20 | ···················· | ···················· | ···················· | ···················· | ···················· | ···················· |
| J | 1832 | hCoV-19/England/MILK-99482E/2020\|EPI_ISL_552291\|20 | ···················· | ···················· | ···················· | ···················· | ···················· | ···················· |
| J | 1833 | hCoV-19/England/MILK-994B61/2020\|EPI_ISL_552293\|20 | ···················· | ···················· | ···················· | ···················· | ·····T·············· | ···················· |
| J | 1834 | hCoV-19/England/MILK-994CE6/2020\|EPI_ISL_552296\|20 | ···················· | ···················· | ···················· | ···················· | ···················· | ···················· |
| J | 1835 | hCoV-19/England/MILK-9946E8/2020\|EPI_ISL_552298\|20 | ···················· | ·A·················· | ···················· | ···················· | ···················· | ··A················· |
| J | 1836 | hCoV-19/England/MILK-994D2F/2020\|EPI_ISL_552299\|20 | ···················· | ···················· | ···················· | ···················· | ···················· | ···················· |
| J | 1837 | hCoV-19/England/MILK-993C7E/2020\|EPI_ISL_552301\|20 | ···················· | ···················· | ···················· | ···················· | ···················· | ···················· |
| J | 1838 | hCoV-19/England/MILK-9947F4/2020\|EPI_ISL_552302\|20 | ···················· | ···················· | ···················· | ···················· | ···················· | ···················· |
| J | 1839 | hCoV-19/England/MILK-993C23/2020\|EPI_ISL_552303\|20 | ···················· | ·A·················· | ···················· | ···················· | ···················· | ···················· |
| J | 1840 | hCoV-19/England/MILK-994CD7/2020\|EPI_ISL_552305\|20 | ···················· | ···················· | ···················· | ···················· | ···················· | ···················· |
| J | 1841 | hCoV-19/England/MILK-993B08/2020\|EPI_ISL_552307\|20 | ···················· | ···················· | ···················· | ···················· | ···················· | ···················· |
| J | 1842 | hCoV-19/England/MILK-99508F/2020\|EPI_ISL_552309\|20 | ···················· | ···················· | ···················· | ···················· | ···················· | ···················· |
| J | 1843 | hCoV-19/England/MILK-994EA4/2020\|EPI_ISL_552310\|20 | ···················· | ·A·················· | ···················· | ···················· | ···················· | ··A················· |
| J | 1844 | hCoV-19/England/MILK-994642/2020\|EPI_ISL_552311\|20 | ···················· | ···················· | ···················· | ···················· | ···················· | ···················· |
| J | 1845 | hCoV-19/England/MILK-994A0A/2020\|EPI_ISL_552312\|20 | ···················· | ···················· | ···················· | ···················· | ···················· | ···················· |
| J | 1846 | hCoV-19/England/MILK-9952A7/2020\|EPI_ISL_552313\|20 | ···················· | ···················· | ···················· | ···················· | ···················· | ···················· |
| J | 1847 | hCoV-19/England/MILK-994CB9/2020\|EPI_ISL_552315\|20 | ···················· | ···················· | ···················· | ···················· | ·····T·············· | ···················· |
| J | 1848 | hCoV-19/England/MILK-994B52/2020\|EPI_ISL_552316\|20 | ···················· | ···················· | ···················· | ···················· | ···················· | ···················· |
| J | 1849 | hCoV-19/England/MILK-994DE3/2020\|EPI_ISL_552317\|20 | ···················· | ···················· | ···················· | ···················· | ···················· | ···················· |
| J | 1850 | hCoV-19/England/MILK-993DB7/2020\|EPI_ISL_552318\|20 | ···················· | ···················· | ···················· | ···················· | ···················· | ···················· |
| J | 1851 | hCoV-19/England/MILK-994A73/2020\|EPI_ISL_552320\|20 | ·······T············ | ···················· | ···················· | ···················· | ···················· | ···················· |
| J | 1852 | hCoV-19/England/MILK-993B53/2020\|EPI_ISL_552321\|20 | ···················· | ···················· | ···················· | ···················· | ···················· | ···················· |
| J | 1853 | hCoV-19/England/MILK-9950BC/2020\|EPI_ISL_552322\|20 | ···················· | ·A·················· | ···················· | ···················· | ···················· | ··A················· |
| J | 1854 | hCoV-19/England/MILK-99386B/2020\|EPI_ISL_552323\|20 | ···················· | ···················· | ···················· | ···················· | ···················· | ···················· |
| J | 1855 | hCoV-19/England/MILK-995359/2020\|EPI_ISL_552324\|20 | ···················· | ···················· | ···················· | ···················· | ···················· | ···················· |
| J | 1856 | hCoV-19/England/MILK-99394A/2020\|EPI_ISL_552325\|20 | ···················· | ···················· | ···················· | ···················· | ···················· | ···················· |
| J | 1857 | hCoV-19/England/MILK-993C9C/2020\|EPI_ISL_552326\|20 | ···················· | ···················· | ···················· | ···················· | ···················· | ···················· |
| J | 1858 | hCoV-19/England/MILK-991CAD/2020\|EPI_ISL_552327\|20 | ···················· | ···················· | ···················· | ···················· | ···················· | ···················· |
| J | 1859 | hCoV-19/England/MILK-99490D/2020\|EPI_ISL_552328\|20 | ···················· | ···················· | ···················· | ···················· | ···················· | ···················· |
| J | 1860 | hCoV-19/England/MILK-994509/2020\|EPI_ISL_552329\|20 | ···················· | ···················· | ···················· | ···················· | ···················· | ···················· |
| J | 1861 | hCoV-19/England/MILK-994E59/2020\|EPI_ISL_552330\|20 | ···················· | ···················· | ···················· | ···················· | ···················· | ···················· |
| J | 1862 | hCoV-19/England/MILK-993A0B/2020\|EPI_ISL_552332\|20 | ···················· | ···················· | ···················· | ···················· | ···················· | ···················· |
| J | 1863 | hCoV-19/Wales/MILK-993A29/2020\|EPI_ISL_552334\|2020 | ···················· | ···················· | ···················· | ···················· | ···················· | ···················· |
| J | 1864 | hCoV-19/England/MILK-99474F/2020\|EPI_ISL_552335\|20 | ···················· | ···················· | ···················· | ···················· | ···················· | ···················· |
| J | 1865 | hCoV-19/England/MILK-9939FF/2020\|EPI_ISL_552336\|20 | ···················· | ···················· | ···················· | ···················· | ···················· | ···················· |
| J | 1866 | hCoV-19/England/MILK-993D6C/2020\|EPI_ISL_552337\|20 | ···················· | ···················· | ···················· | ···················· | ···················· | ···················· |
| J | 1867 | hCoV-19/England/MILK-993C6F/2020\|EPI_ISL_552338\|20 | ···················· | ···················· | ···················· | ···················· | ···················· | ···················· |
| J | 1868 | hCoV-19/England/MILK-9951B9/2020\|EPI_ISL_552339\|20 | ···················· | ·A·················· | ···················· | ···················· | ···················· | ··A················· |
| J | 1869 | hCoV-19/England/MILK-995201/2020\|EPI_ISL_552340\|20 | ···················· | ···················· | ···················· | ···················· | ···················· | ···················· |
| J | 1870 | hCoV-19/England/MILK-994B25/2020\|EPI_ISL_552341\|20 | ···················· | ···················· | ···················· | ···················· | ···················· | ···················· |
| J | 1871 | hCoV-19/England/MILK-995043/2020\|EPI_ISL_552343\|20 | ···················· | ···················· | ···················· | ···················· | ·····T·············· | ···················· |
| J | 1872 | hCoV-19/England/MILK-993D7B/2020\|EPI_ISL_552344\|20 | ···················· | ···················· | ···················· | ···················· | ···················· | ···················· |
| J | 1873 | hCoV-19/England/MILK-995289/2020\|EPI_ISL_552345\|20 | ···················· | ···················· | ···················· | ···················· | ···················· | ···················· |
| J | 1874 | hCoV-19/England/MILK-9944C0/2020\|EPI_ISL_552346\|20 | ···················· | ···················· | ···················· | ···················· | ···················· | ···················· |
| J | 1875 | hCoV-19/England/MILK-9954FC/2020\|EPI_ISL_552347\|20 | ···················· | ···················· | ···················· | ···················· | ···················· | ···················· |
| J | 1876 | hCoV-19/England/MILK-993CBA/2020\|EPI_ISL_552348\|20 | ···················· | ···················· | ···················· | ···················· | ···················· | ···················· |
| J | 1877 | hCoV-19/England/MILK-994ED1/2020\|EPI_ISL_552349\|20 | ···················· | ·A·················· | ···················· | ···················· | ···················· | ··A················· |
| J | 1878 | hCoV-19/England/MILK-995061/2020\|EPI_ISL_552350\|20 | ···················· | ···················· | ···················· | ···················· | ···················· | ···················· |
| J | 1879 | hCoV-19/England/MILK-994E0E/2020\|EPI_ISL_552351\|20 | ···················· | ·A·················· | ···················· | ···················· | ···················· | ··A················· |
| J | 1880 | hCoV-19/England/MILK-994E3B/2020\|EPI_ISL_552352\|20 | ···················· | ···················· | ···················· | ···················· | ···················· | ···················· |
| J | 1881 | hCoV-19/England/MILK-9946BB/2020\|EPI_ISL_552353\|20 | ···················· | ·A·················· | ···················· | ···················· | ···················· | ···················· |
| J | 1882 | hCoV-19/England/MILK-993801/2020\|EPI_ISL_552354\|20 | ···················· | ···················· | ···················· | ···················· | ···················· | ···················· |
| J | 1883 | hCoV-19/England/MILK-993B62/2020\|EPI_ISL_552355\|20 | ···················· | ···················· | ···················· | ···················· | ···················· | ···················· |
| J | 1884 | hCoV-19/England/ALDP-95CA4A/2020\|EPI_ISL_553599\|20 | ···················· | ···················· | ···················· | ···················· | ···················· | ···················· |
| J | 1885 | hCoV-19/England/ALDP-952D96/2020\|EPI_ISL_553632\|20 | ···················· | ···················· | ···················· | ···················· | ···················· | ···················· |
| J | 1886 | hCoV-19/England/ALDP-9530D8/2020\|EPI_ISL_553633\|20 | ···················· | ·A·················· | ···················· | ···················· | ···················· | ···················· |
| J | 1887 | hCoV-19/England/ALDP-952DD2/2020\|EPI_ISL_553634\|20 | ···················· | ···················· | ···················· | ···················· | ···················· | ···················· |
| J | 1888 | hCoV-19/England/ALDP-953919/2020\|EPI_ISL_553635\|20 | ···················· | ···················· | ···················· | ···················· | ···················· | ···················· |
| J | 1889 | hCoV-19/England/ALDP-9521A9/2020\|EPI_ISL_553636\|20 | ···················· | ···················· | ···················· | ···················· | ···················· | ···················· |
| J | 1890 | hCoV-19/England/ALDP-95308D/2020\|EPI_ISL_553637\|20 | ···················· | ···················· | ···················· | ···················· | ···················· | ···················· |
| J | 1891 | hCoV-19/England/ALDP-952E39/2020\|EPI_ISL_553638\|20 | ···················· | ···················· | ···················· | ···················· | ···················· | ···················· |
| J | 1892 | hCoV-19/England/ALDP-95219A/2020\|EPI_ISL_553639\|20 | ···················· | ···················· | ···················· | ···················· | ···················· | ···················· |
| J | 1893 | hCoV-19/England/ALDP-95207F/2020\|EPI_ISL_553641\|20 | ···················· | ·A·················· | ···················· | ···················· | ···················· | ···················· |
| J | 1894 | hCoV-19/England/ALDP-95218B/2020\|EPI_ISL_553642\|20 | ···················· | ···················· | ···················· | ···················· | ···················· | ···················· |
| J | 1895 | hCoV-19/England/ALDP-95222E/2020\|EPI_ISL_553643\|20 | ···················· | ···················· | ···················· | ···················· | ···················· | ···················· |

| | | | | | | | | |
|---|---|---|---|---|---|---|---|---|
| J | 1896 | hCoV-19/England/ALDP-9538FE/2020\|EPI_ISL_553644\|20 | ···················· | ·A·················· | ···················· | ···················· | ···················· | ···················· |
| J | 1897 | hCoV-19/England/ALDP-952E57/2020\|EPI_ISL_553645\|20 | ···················· | ···················· | ···················· | ···················· | ···················· | ···················· |
| J | 1898 | hCoV-19/England/ALDP-95221F/2020\|EPI_ISL_553646\|20 | ···················· | ···················· | ···················· | ···················· | ···················· | ···················· |
| J | 1899 | hCoV-19/England/ALDP-952358/2020\|EPI_ISL_553647\|20 | ···················· | ···················· | ···················· | ···················· | ···················· | ···················· |
| J | 1900 | hCoV-19/England/ALDP-9520D9/2020\|EPI_ISL_553651\|20 | ···················· | ···················· | ···················· | ···················· | ···················· | ···················· |
| J | 1901 | hCoV-19/England/ALDP-952E93/2020\|EPI_ISL_553653\|20 | ···················· | ···················· | ···················· | ···················· | ···················· | ···················· |
| J | 1902 | hCoV-19/England/ALDP-952E2A/2020\|EPI_ISL_553655\|20 | ···················· | ···················· | ···················· | ···················· | ···················· | ···················· |
| J | 1903 | hCoV-19/England/ALDP-952200/2020\|EPI_ISL_553656\|20 | ···················· | ···················· | ···················· | ···················· | ···················· | ···················· |
| J | 1904 | hCoV-19/England/ALDP-952DC3/2020\|EPI_ISL_553657\|20 | ···················· | ···················· | ···················· | ···················· | ···················· | ···················· |
| J | 1905 | hCoV-19/England/ALDP-9521D6/2020\|EPI_ISL_553660\|20 | ···················· | ···················· | ···················· | ···················· | ···················· | ···················· |
| J | 1906 | hCoV-19/England/ALDP-952D69/2020\|EPI_ISL_553661\|20 | ···················· | ···················· | ···················· | ···················· | ···················· | ···················· |
| J | 1907 | hCoV-19/England/ALDP-95223D/2020\|EPI_ISL_553662\|20 | ···················· | ···················· | ···················· | ···················· | ···················· | ···················· |
| J | 1908 | hCoV-19/England/ALDP-953014/2020\|EPI_ISL_553663\|20 | ···················· | ···················· | ···················· | ···················· | ···················· | ···················· |
| J | 1909 | hCoV-19/England/ALDP-9521E5/2020\|EPI_ISL_553666\|20 | ···················· | ·A·················· | ···················· | ···················· | ···················· | ···················· |
| J | 1910 | hCoV-19/England/ALDP-952EEE/2020\|EPI_ISL_553668\|20 | ···················· | ···················· | ···················· | ···················· | ···················· | ···················· |
| J | 1911 | hCoV-19/England/ALDP-9530AB/2020\|EPI_ISL_553669\|20 | ···················· | ···················· | ···················· | ···················· | ···················· | ···················· |
| J | 1912 | hCoV-19/England/ALDP-952F72/2020\|EPI_ISL_553670\|20 | ···················· | ···················· | ···················· | ···················· | ···················· | ···················· |
| J | 1913 | hCoV-19/England/ALDP-952F36/2020\|EPI_ISL_553671\|20 | ···················· | ···················· | ···················· | ···················· | ···················· | ···················· |
| J | 1914 | hCoV-19/England/ALDP-952F81/2020\|EPI_ISL_553672\|20 | ···················· | ···················· | ···················· | ···················· | ···················· | ···················· |
| J | 1915 | hCoV-19/England/ALDP-952EDF/2020\|EPI_ISL_553673\|20 | ···················· | ···················· | ···················· | ···················· | ···················· | ···················· |
| J | 1916 | hCoV-19/England/ALDP-9521F4/2020\|EPI_ISL_553675\|20 | ···················· | ···················· | ···················· | ···················· | ···················· | ···················· |
| J | 1917 | hCoV-19/England/ALDP-9521C7/2020\|EPI_ISL_553676\|20 | ···················· | ···················· | ···················· | ···················· | ···················· | ···················· |
| J | 1918 | hCoV-19/England/ALDP-952FFA/2020\|EPI_ISL_553677\|20 | ···················· | ···················· | ···················· | ···················· | ···················· | ···················· |
| J | 1919 | hCoV-19/England/ALDP-953050/2020\|EPI_ISL_553678\|20 | ···················· | ···················· | ···················· | ···················· | ···················· | ···················· |
| J | 1920 | hCoV-19/England/ALDP-952E66/2020\|EPI_ISL_553679\|20 | ···················· | ···················· | ···················· | ···················· | ···················· | ···················· |
| J | 1921 | hCoV-19/England/ALDP-952F27/2020\|EPI_ISL_553680\|20 | ···················· | ···················· | ···················· | ···················· | ···················· | ···················· |
| J | 1922 | hCoV-19/England/ALDP-953111/2020\|EPI_ISL_553681\|20 | ···················· | ···················· | ···················· | ···················· | ···················· | ···················· |
| J | 1923 | hCoV-19/England/ALDP-9520CA/2020\|EPI_ISL_553683\|20 | ···················· | ···················· | ···················· | ···················· | ···················· | ···················· |
| J | 1924 | hCoV-19/England/ALDP-95309C/2020\|EPI_ISL_553685\|20 | ···················· | ···················· | ···················· | ···················· | ···················· | ···················· |
| J | 1925 | hCoV-19/England/ALDP-952E48/2020\|EPI_ISL_553686\|20 | ···················· | ···················· | ···················· | ···················· | ···················· | ···················· |
| J | 1926 | hCoV-19/England/ALDP-94D7EB/2020\|EPI_ISL_554963\|20 | ···················· | ···················· | ···················· | ···················· | ···················· | ···················· |
| J | 1927 | hCoV-19/England/ALDP-95A9B8/2020\|EPI_ISL_554967\|20 | ···················· | ···················· | ···················· | ···················· | ···················· | ···················· |
| J | 1928 | hCoV-19/England/ALDP-959EBA/2020\|EPI_ISL_554968\|20 | ···················· | ···················· | ···················· | ···················· | ···················· | ···················· |
| J | 1929 | hCoV-19/England/ALDP-95CA95/2020\|EPI_ISL_554971\|20 | ···················· | ·A·················· | ···················· | ···················· | ···················· | ···················· |
| J | 1930 | hCoV-19/Wales/ALDP-95C2CA/2020\|EPI_ISL_554972\|2020 | ···················· | ···················· | ···················· | ···················· | ···················· | ···················· |
| J | 1931 | hCoV-19/England/ALDP-94D824/2020\|EPI_ISL_554973\|20 | ···················· | ···················· | ···················· | ···················· | ···················· | ···················· |
| J | 1932 | hCoV-19/England/ALDP-95CAA4/2020\|EPI_ISL_554974\|20 | ···················· | ···················· | ···················· | ···················· | ···················· | ···················· |
| J | 1933 | hCoV-19/England/ALDP-95C127/2020\|EPI_ISL_554975\|20 | ···················· | ···················· | ···················· | ···················· | ···················· | ···················· |
| J | 1934 | hCoV-19/Wales/ALDP-94D781/2020\|EPI_ISL_554976\|2020 | ···················· | ···················· | ···················· | ···················· | ···················· | ···················· |
| J | 1935 | hCoV-19/England/ALDP-95CD14/2020\|EPI_ISL_554977\|20 | ···················· | ···················· | ···················· | ···················· | ···················· | ···················· |
| J | 1936 | hCoV-19/England/ALDP-95A96D/2020\|EPI_ISL_554981\|20 | ···················· | ···················· | ···················· | ···················· | ···················· | ···················· |
| J | 1937 | hCoV-19/England/ALDP-95A9A9/2020\|EPI_ISL_554982\|20 | ···················· | ·A·················· | ···················· | ···················· | ···················· | ···················· |
| J | 1938 | hCoV-19/England/ALDP-95C646/2020\|EPI_ISL_554985\|20 | ···················· | ···················· | ···················· | ···················· | ···················· | ···················· |
| J | 1939 | hCoV-19/England/ALDP-95CA86/2020\|EPI_ISL_554986\|20 | ···················· | ···················· | ···················· | ···················· | ···················· | ···················· |
| J | 1940 | hCoV-19/England/ALDP-95C2AC/2020\|EPI_ISL_554988\|20 | ···················· | ···················· | ···················· | ···················· | ···················· | ···················· |
| J | 1941 | hCoV-19/England/ALDP-95D029/2020\|EPI_ISL_554989\|20 | ···················· | ···················· | ···················· | ···················· | ···················· | ···················· |
| J | 1942 | hCoV-19/England/ALDP-95CE11/2020\|EPI_ISL_554991\|20 | ···················· | ···················· | ···················· | ···················· | ···················· | ···················· |
| J | 1943 | hCoV-19/England/ALDP-95CDE7/2020\|EPI_ISL_554992\|20 | ···················· | ···················· | ···················· | ···················· | ···················· | ···················· |
| J | 1944 | hCoV-19/Wales/ALDP-94D790/2020\|EPI_ISL_554993\|2020 | ···················· | ···················· | ···················· | ···················· | ···················· | ···················· |
| J | 1945 | hCoV-19/England/ALDP-95AB0D/2020\|EPI_ISL_554994\|20 | ···················· | ···················· | ···················· | ···················· | ···················· | ···················· |
| J | 1946 | hCoV-19/England/ALDP-94D51E/2020\|EPI_ISL_554995\|20 | ···················· | ···················· | ···················· | ···················· | ···················· | ···················· |
| J | 1947 | hCoV-19/England/ALDP-95A9D6/2020\|EPI_ISL_554997\|20 | ···················· | ···················· | ···················· | ···················· | ···················· | ···················· |
| J | 1948 | hCoV-19/England/ALDP-95C682/2020\|EPI_ISL_554998\|20 | ···················· | ···················· | ···················· | ···················· | ···················· | ···················· |
| J | 1949 | hCoV-19/England/ALDP-95CD05/2020\|EPI_ISL_554999\|20 | ···················· | ···················· | ···················· | ···················· | ···················· | ···················· |
| J | 1950 | hCoV-19/England/ALDP-8FFCF9/2020\|EPI_ISL_557130\|20 | ···················· | ···················· | ···················· | ···················· | ···················· | ···················· |
| J | 1951 | hCoV-19/England/ALDP-8FF8C8/2020\|EPI_ISL_557131\|20 | ···················· | ···················· | ···················· | ···················· | ···················· | ···················· |
| J | 1952 | hCoV-19/England/ALDP-8B5E35/2020\|EPI_ISL_557133\|20 | ···················· | ···················· | ···················· | ···················· | ···················· | ···················· |
| J | 1953 | hCoV-19/England/ALDP-8E2C53/2020\|EPI_ISL_557135\|20 | ···················· | ···················· | ···················· | ···················· | ···················· | ···················· |
| J | 1954 | hCoV-19/England/ALDP-8D1255/2020\|EPI_ISL_557136\|20 | ···················· | ···················· | ···················· | ···················· | ···················· | ···················· |
| J | 1955 | hCoV-19/England/ALDP-90DAA2/2020\|EPI_ISL_557137\|20 | ···················· | ···················· | ···················· | ···················· | ···················· | ···················· |
| J | 1956 | hCoV-19/England/ALDP-8B9FD4/2020\|EPI_ISL_557138\|20 | ···················· | ···················· | ···················· | ···················· | ···················· | ···················· |
| J | 1957 | hCoV-19/England/ALDP-8CF1A8/2020\|EPI_ISL_557139\|20 | ···················· | ···················· | ···················· | ···················· | ···················· | ···················· |
| J | 1958 | hCoV-19/England/ALDP-8FDE22/2020\|EPI_ISL_557141\|20 | ···················· | ···················· | ···················· | ···················· | ···················· | ···················· |
| J | 1959 | hCoV-19/England/ALDP-8E25EF/2020\|EPI_ISL_557143\|20 | ···················· | ···················· | ···················· | ···················· | ···················· | ···················· |
| J | 1960 | hCoV-19/England/ALDP-8D4771/2020\|EPI_ISL_557144\|20 | ···················· | ···················· | ···················· | ···················· | ···················· | ···················· |
| J | 1961 | hCoV-19/England/ALDP-8B9402/2020\|EPI_ISL_557145\|20 | ···················· | ···················· | ···················· | ···················· | ···················· | ···················· |
| J | 1962 | hCoV-19/England/ALDP-90DB90/2020\|EPI_ISL_557147\|20 | ···················· | ···················· | ···················· | ···················· | ···················· | ···················· |
| J | 1963 | hCoV-19/England/ALDP-8DE249/2020\|EPI_ISL_557148\|20 | ···················· | ···················· | ···················· | ···················· | ···················· | ···················· |
| J | 1964 | hCoV-19/England/ALDP-8B53EB/2020\|EPI_ISL_557150\|20 | ···················· | ···················· | ···················· | ···················· | ···················· | ···················· |
| J | 1965 | hCoV-19/England/ALDP-8D5190/2020\|EPI_ISL_557151\|20 | ···················· | ···················· | ···················· | ···················· | ···················· | ···················· |
| J | 1966 | hCoV-19/England/ALDP-90FB70/2020\|EPI_ISL_557152\|20 | ···················· | ···················· | ···················· | ···················· | ···················· | ···················· |
| J | 1967 | hCoV-19/England/ALDP-8B65F3/2020\|EPI_ISL_557153\|20 | ···················· | ···················· | ···················· | ···················· | ···················· | ···················· |
| J | 1968 | hCoV-19/England/ALDP-9014EC/2020\|EPI_ISL_557154\|20 | ···················· | ···················· | ···················· | ···················· | ···················· | ···················· |
| J | 1969 | hCoV-19/England/ALDP-8CF849/2020\|EPI_ISL_557155\|20 | ···················· | ···················· | ···················· | ···················· | ···················· | ···················· |
| J | 1970 | hCoV-19/England/ALDP-8DF7B2/2020\|EPI_ISL_557157\|20 | ···················· | ···················· | ···················· | ···················· | ···················· | ···················· |
| J | 1971 | hCoV-19/England/ALDP-8D640F/2020\|EPI_ISL_557158\|20 | ···················· | ···················· | ···················· | ···················· | ···················· | ···················· |
| J | 1972 | hCoV-19/England/ALDP-8E3988/2020\|EPI_ISL_557159\|20 | ···················· | ···················· | ···················· | ···················· | ···················· | ···················· |
| J | 1973 | hCoV-19/England/MILK-594FF0/2020\|EPI_ISL_558400\|20 | ···················· | ···················· | ···················· | ···················· | ···················· | ···················· |
| J | 1974 | hCoV-19/England/MILK-57A840/2020\|EPI_ISL_558401\|20 | ···················· | ···················· | ···················· | ···················· | ···················· | ···················· |
| J | 1975 | hCoV-19/England/ALDP-5D28FB/2020\|EPI_ISL_558404\|20 | ···················· | ···················· | ···················· | ···················· | ···················· | ···················· |

| | | | | | | | | |
|---|---|---|---|---|---|---|---|---|
| J | 1976 | hCoV-19/England/ALDP-5D009B/2020\|EPI_ISL_558450\|20 | ···················· | ···················· | ···················· | ···················· | ···················· | ···················· |
| J | 1977 | hCoV-19/England/ALDP-5CFE86/2020\|EPI_ISL_558451\|20 | ···················· | ···················· | ···················· | ···················· | ···················· | ···················· |
| J | 1978 | hCoV-19/England/ALDP-5CB215/2020\|EPI_ISL_558453\|20 | ···················· | ···················· | ···················· | ···················· | ···················· | ···················· |
| J | 1979 | hCoV-19/England/ALDP-5D02FF/2020\|EPI_ISL_558454\|20 | ···················· | ···················· | ···················· | ···················· | ···················· | ···················· |
| J | 1980 | hCoV-19/England/ALDP-5D63BA/2020\|EPI_ISL_558455\|20 | ···················· | ···················· | ···················· | ···················· | ···················· | ···················· |
| J | 1981 | hCoV-19/England/ALDP-5CF448/2020\|EPI_ISL_558457\|20 | ···················· | ···················· | ···················· | ···················· | ···················· | ···················· |
| J | 1982 | hCoV-19/England/ALDP-5C6599/2020\|EPI_ISL_558458\|20 | ···················· | ·············A······ | ···················· | ···················· | ···················· | ···················· |
| J | 1983 | hCoV-19/England/ALDP-5CC2F6/2020\|EPI_ISL_558459\|20 | ···················· | ···················· | ···················· | ···················· | ···················· | ···················· |
| J | 1984 | hCoV-19/England/ALDP-5CF536/2020\|EPI_ISL_558462\|20 | ···················· | ···················· | ···················· | ···················· | ···················· | ···················· |
| J | 1985 | hCoV-19/England/ALDP-5D3933/2020\|EPI_ISL_558463\|20 | ···················· | ···················· | ···················· | ···················· | ···················· | ···················· |
| J | 1986 | hCoV-19/England/ALDP-5CDC9D/2020\|EPI_ISL_558464\|20 | ···················· | ···················· | ···················· | ···················· | ···················· | ···················· |
| J | 1987 | hCoV-19/England/ALDP-5D52BE/2020\|EPI_ISL_558465\|20 | ···················· | ···················· | ···················· | ···················· | ···················· | ···················· |
| J | 1988 | hCoV-19/England/ALDP-5D2A04/2020\|EPI_ISL_558467\|20 | ···················· | ···················· | ···················· | ···················· | ···················· | ···················· |
| J | 1989 | hCoV-19/England/ALDP-5CBAE0/2020\|EPI_ISL_558468\|20 | ···················· | ···················· | ···················· | ···················· | ···················· | ···················· |
| J | 1990 | hCoV-19/England/ALDP-5C88AD/2020\|EPI_ISL_558469\|20 | ···················· | ···················· | ···················· | ···················· | ···················· | ···················· |
| J | 1991 | hCoV-19/England/ALDP-5D067B/2020\|EPI_ISL_558470\|20 | ···················· | ···················· | ···················· | ···················· | ···················· | ···················· |
| J | 1992 | hCoV-19/England/ALDP-5D9D40/2020\|EPI_ISL_558472\|20 | ···················· | ···················· | ···················· | ···················· | ···················· | ···················· |
| J | 1993 | hCoV-19/England/ALDP-5CE1BB/2020\|EPI_ISL_558473\|20 | ···················· | ···················· | ···················· | ···················· | ···················· | ···················· |
| J | 1994 | hCoV-19/England/ALDP-5D149E/2020\|EPI_ISL_558475\|20 | ···················· | ···················· | ···················· | ···················· | ···················· | ···················· |
| J | 1995 | hCoV-19/England/ALDP-5D92AB/2020\|EPI_ISL_558476\|20 | ···················· | ···················· | ···················· | ···················· | ···················· | ···················· |
| J | 1996 | hCoV-19/England/ALDP-5CB36D/2020\|EPI_ISL_558477\|20 | ···················· | ···················· | ···················· | ···················· | ···················· | ···················· |
| J | 1997 | hCoV-19/England/ALDP-5C789F/2020\|EPI_ISL_558478\|20 | ···················· | ···················· | ···················· | ···················· | ···················· | ···················· |
| J | 1998 | hCoV-19/England/ALDP-5C96EE/2020\|EPI_ISL_558479\|20 | ···················· | ···················· | ···················· | ···················· | ···················· | ···················· |
| J | 1999 | hCoV-19/England/ALDP-5D1461/2020\|EPI_ISL_558480\|20 | ···················· | ···················· | ···················· | ···················· | ···················· | ···················· |
| J | 2000 | hCoV-19/England/ALDP-5C70C4/2020\|EPI_ISL_558482\|20 | ···················· | ···················· | ···················· | ···················· | ···················· | ···················· |
| J | 2001 | hCoV-19/England/ALDP-5D78BA/2020\|EPI_ISL_558483\|20 | ···················· | ···················· | ···················· | ···················· | ···················· | ···················· |
| J | 2002 | hCoV-19/England/ALDP-5CD1DA/2020\|EPI_ISL_558484\|20 | ···················· | ···················· | ···················· | ···················· | ···················· | ···················· |
| J | 2003 | hCoV-19/England/ALDP-5D9A0D/2020\|EPI_ISL_558486\|20 | ···················· | ···················· | ···················· | ···················· | ···················· | ···················· |
| J | 2004 | hCoV-19/England/ALDP-5CB38B/2020\|EPI_ISL_558488\|20 | ···················· | ···················· | ···················· | ···················· | ···················· | ···················· |
| J | 2005 | hCoV-19/England/ALDP-5D1CA5/2020\|EPI_ISL_558489\|20 | ···················· | ···················· | ···················· | ···················· | ···················· | ···················· |
| J | 2006 | hCoV-19/England/MILK-4858F8/2020\|EPI_ISL_559701\|20 | ···················· | ···················· | ···················· | ···················· | ···················· | ···················· |
| J | 2007 | hCoV-19/England/MILK-4868E8/2020\|EPI_ISL_559702\|20 | ···················· | ···················· | ···················· | ···················· | ···················· | ···················· |
| J | 2008 | hCoV-19/England/MILK-46BBC2/2020\|EPI_ISL_559703\|20 | ···················· | ···················· | ···················· | ···················· | ···················· | ···················· |
| J | 2009 | hCoV-19/England/MILK-45F6DF/2020\|EPI_ISL_559704\|20 | ···················· | ···················· | ···················· | ···················· | ···················· | ···················· |
| J | 2010 | hCoV-19/England/MILK-4619FF/2020\|EPI_ISL_559705\|20 | ···················· | ···················· | ···················· | ···················· | ···················· | ···················· |
| J | 2011 | hCoV-19/England/MILK-460B27/2020\|EPI_ISL_559707\|20 | ···················· | ···················· | ···················· | ···················· | ···················· | ···················· |
| J | 2012 | hCoV-19/England/MILK-45C21A/2020\|EPI_ISL_559708\|20 | ···················· | ···················· | ···················· | ···················· | ···················· | ···················· |
| J | 2013 | hCoV-19/England/MILK-46AAC6/2020\|EPI_ISL_559710\|20 | ···················· | ···················· | ···················· | ···················· | ···················· | ···················· |
| J | 2014 | hCoV-19/England/MILK-46D2F8/2020\|EPI_ISL_559711\|20 | ···················· | ···················· | ···················· | ···················· | ···················· | ···················· |
| J | 2015 | hCoV-19/England/MILK-4675F5/2020\|EPI_ISL_559712\|20 | ···················· | ···················· | ···················· | ···················· | ···················· | ···················· |
| J | 2016 | hCoV-19/England/MILK-45ED08/2020\|EPI_ISL_559713\|20 | ···················· | ···················· | ···················· | ···················· | ···················· | ···················· |
| J | 2017 | hCoV-19/England/MILK-4699F7/2020\|EPI_ISL_559714\|20 | ···················· | ···················· | ···················· | ···················· | ···················· | ···················· |
| J | 2018 | hCoV-19/England/MILK-45CE8F/2020\|EPI_ISL_559716\|20 | ···················· | ···················· | ··Y················· | ···········W········ | ···················· | ···················· |
| J | 2019 | hCoV-19/England/MILK-46804E/2020\|EPI_ISL_559718\|20 | ···················· | ···················· | ···················· | ···················· | ···················· | ···················· |
| J | 2020 | hCoV-19/England/MILK-4674DA/2020\|EPI_ISL_559719\|20 | ···················· | ···················· | ···················· | ···················· | ···················· | ···················· |
| J | 2021 | hCoV-19/England/MILK-46822A/2020\|EPI_ISL_559720\|20 | ···················· | ···················· | ···················· | ···················· | ···················· | ···················· |
| J | 2022 | hCoV-19/England/MILK-468D74/2020\|EPI_ISL_559721\|20 | ···················· | ···················· | ···················· | ···················· | ···················· | ···················· |
| J | 2023 | hCoV-19/England/MILK-46C00E/2020\|EPI_ISL_559722\|20 | ···················· | ···················· | ···················· | ···················· | ···················· | ···················· |
| J | 2024 | hCoV-19/England/MILK-467434/2020\|EPI_ISL_559723\|20 | ···················· | ···················· | ···················· | ···················· | ···················· | ···················· |
| J | 2025 | hCoV-19/England/MILK-469766/2020\|EPI_ISL_559725\|20 | ···················· | ···················· | ···················· | ···················· | ···················· | ···················· |
| J | 2026 | hCoV-19/England/MILK-467F7F/2020\|EPI_ISL_559726\|20 | ···················· | ···················· | ···················· | ···················· | ···················· | ···················· |
| J | 2027 | hCoV-19/England/MILK-469924/2020\|EPI_ISL_559727\|20 | ···················· | ···················· | ···················· | ···················· | ···················· | ···················· |
| J | 2028 | hCoV-19/England/MILK-461A74/2020\|EPI_ISL_559728\|20 | ···················· | ·A·················· | ···················· | ···················· | ···················· | ···················· |
| J | 2029 | hCoV-19/England/MILK-4673CE/2020\|EPI_ISL_559729\|20 | ···················· | ···················· | ···················· | ···················· | ···················· | ···················· |
| J | 2030 | hCoV-19/England/MILK-45F7EB/2020\|EPI_ISL_559730\|20 | ···················· | ·R·················· | ···················· | ···················· | ···················· | ···················· |
| J | 2031 | hCoV-19/England/MILK-467DEE/2020\|EPI_ISL_559731\|20 | ···················· | ···················· | ···················· | ···················· | ···················· | ···················· |
| J | 2032 | hCoV-19/England/MILK-45DD72/2020\|EPI_ISL_559732\|20 | ···················· | ···················· | ··Y················· | ···········W········ | ···················· | ···················· |
| J | 2033 | hCoV-19/England/MILK-46A3E9/2020\|EPI_ISL_559735\|20 | ···················· | ···················· | ···················· | ···················· | ···················· | ···················· |
| J | 2034 | hCoV-19/England/MILK-45E61C/2020\|EPI_ISL_559736\|20 | ···················· | ···················· | ···················· | ···················· | ···················· | ···················· |
| J | 2035 | hCoV-19/England/QEUH-99C02E/2020\|EPI_ISL_559780\|20 | ···················· | ···················· | ···················· | ···················· | ···················· | ···················· |
| J | 2036 | hCoV-19/England/QEUH-99C04C/2020\|EPI_ISL_559781\|20 | ···················· | ···················· | ···················· | ···················· | ···················· | ···················· |
| J | 2037 | hCoV-19/England/QEUH-99C097/2020\|EPI_ISL_559782\|20 | ···················· | ···················· | ···················· | ···················· | ···················· | ···················· |
| J | 2038 | hCoV-19/England/QEUH-99C0A6/2020\|EPI_ISL_559783\|20 | ···················· | ···················· | ···················· | ···················· | ···················· | ···················· |
| J | 2039 | hCoV-19/England/QEUH-99C10D/2020\|EPI_ISL_559784\|20 | ···················· | ···················· | ···················· | ···················· | ···················· | ···················· |
| J | 2040 | hCoV-19/England/QEUH-99C167/2020\|EPI_ISL_559786\|20 | ···················· | ···················· | ···················· | ···················· | ···················· | ···················· |
| J | 2041 | hCoV-19/England/QEUH-99C1B2/2020\|EPI_ISL_559787\|20 | ···················· | ···················· | ···················· | ···················· | ···················· | ···················· |
| J | 2042 | hCoV-19/England/QEUH-99C1C1/2020\|EPI_ISL_559788\|20 | ···················· | ···················· | ···················· | ···················· | ···················· | ···················· |
| J | 2043 | hCoV-19/England/QEUH-99C1FE/2020\|EPI_ISL_559789\|20 | ···················· | ···················· | ···················· | ···················· | ······T············· | ···················· |
| J | 2044 | hCoV-19/England/QEUH-99C325/2020\|EPI_ISL_559793\|20 | ···················· | ·A·················· | ···················· | ···················· | ···················· | ···A················ |
| J | 2045 | hCoV-19/England/QEUH-99C361/2020\|EPI_ISL_559794\|20 | ···················· | ···················· | ···················· | ···················· | ···················· | ···················· |
| J | 2046 | hCoV-19/England/QEUH-99C39E/2020\|EPI_ISL_559796\|20 | ···················· | ···················· | ···················· | ···················· | ···················· | ···················· |
| J | 2047 | hCoV-19/England/QEUH-99C3AD/2020\|EPI_ISL_559797\|20 | ···················· | ···················· | ···················· | ···················· | ···················· | ···················· |
| J | 2048 | hCoV-19/England/QEUH-9ACB93/2020\|EPI_ISL_559799\|20 | ···················· | ···················· | ···················· | ···················· | ···················· | ···················· |
| J | 2049 | hCoV-19/England/CAMC-9B1EB0/2020\|EPI_ISL_549400\|20 | ···················· | ···················· | ···················· | ···················· | ······T············· | ···················· |
| J | 2050 | hCoV-19/England/CAMC-9B1EFC/2020\|EPI_ISL_549401\|20 | ···················· | ···················· | ···················· | ···················· | ···················· | ···················· |
| J | 2051 | hCoV-19/England/CAMC-9B1F08/2020\|EPI_ISL_549402\|20 | ···················· | ···················· | ···················· | ···················· | ···················· | ···················· |
| J | 2052 | hCoV-19/England/CAMC-9B1F26/2020\|EPI_ISL_549403\|20 | ···················· | ···················· | ···················· | ···················· | ···················· | ···················· |
| J | 2053 | hCoV-19/England/CAMC-9B1FEA/2020\|EPI_ISL_549404\|20 | ···················· | ···················· | ···················· | ···················· | ···················· | ···················· |
| J | 2054 | hCoV-19/England/CAMC-9B2004/2020\|EPI_ISL_549405\|20 | ···················· | ···················· | ···················· | ···················· | ···················· | ···················· |
| J | 2055 | hCoV-19/England/CAMC-9B22D1/2020\|EPI_ISL_549406\|20 | ···················· | ···················· | ···················· | ···················· | ···················· | ···················· |

J 2056 hCoV-19/England/CAMC-9B22FF/2020|EPI_ISL_549407|20 ···················· ···················· ····················· ···················· ···················· ····················
J 2057 hCoV-19/England/CAMC-9B2462/2020|EPI_ISL_549410|20 ···················· ···················· ····················· ···················· ···················· ····················
J 2058 hCoV-19/England/CAMC-9B24F9/2020|EPI_ISL_549411|20 ···················· ···················· ····················· ···················· ···················· ····················
J 2059 hCoV-19/England/CAMC-9B2514/2020|EPI_ISL_549412|20 ···················· ···················· ····················· ···················· ···················· ····················
J 2060 hCoV-19/England/CAMC-9B29EB/2020|EPI_ISL_549413|20 ···················· ···················· ····················· ···················· ···················· ····················
J 2061 hCoV-19/England/MILK-9A86C3/2020|EPI_ISL_549414|20 ···················· ···················· ····················· ···················· ···················· ····················
J 2062 hCoV-19/England/MILK-9A8757/2020|EPI_ISL_549417|20 ···················· ···················· ····················· ···················· ···················· ····················
J 2063 hCoV-19/England/MILK-9A8766/2020|EPI_ISL_549418|20 ···················· ·A·················· ····················· ···················· ···················· ····················
J 2064 hCoV-19/England/MILK-9A96A4/2020|EPI_ISL_551025|20 ···················· ···················· ····················· ···················· ···················· ····················
J 2065 hCoV-19/England/MILK-9AB253/2020|EPI_ISL_551026|20 ···················· ···················· ····················· ···················· ···················· ····················
J 2066 hCoV-19/England/MILK-9A962C/2020|EPI_ISL_551027|20 ···················· ···················· ····················· ···················· ···················· ····················
J 2067 hCoV-19/England/MILK-9A8FD7/2020|EPI_ISL_551028|20 ···················· ···················· ····················· ···················· ···················· ····················
J 2068 hCoV-19/England/MILK-9A9B1E/2020|EPI_ISL_551029|20 ···················· ···················· ····················· ···················· ·····T·············· ····················
J 2069 hCoV-19/England/MILK-9A97B0/2020|EPI_ISL_551030|20 ···················· ···················· ····················· ···················· ···················· ····················
J 2070 hCoV-19/England/MILK-9AB43F/2020|EPI_ISL_551031|20 ···················· ···················· ····················· ···················· ···················· ····················
J 2071 hCoV-19/England/MILK-9AB48A/2020|EPI_ISL_551032|20 ···················· ···················· ····················· ···················· ···················· ····················
J 2072 hCoV-19/England/MILK-9A954D/2020|EPI_ISL_551034|20 ···················· ···················· ····················· ···················· ···················· ····················
J 2073 hCoV-19/England/MILK-9A94D7/2020|EPI_ISL_551035|20 ···················· ···················· ····················· ···················· ···················· ····················
J 2074 hCoV-19/England/MILK-9A946E/2020|EPI_ISL_551036|20 ···················· ···················· ····················· ···················· ···················· ····················
J 2075 hCoV-19/England/MILK-9A90E2/2020|EPI_ISL_551037|20 ···················· ···················· ····················· ···················· ···················· ····················
J 2076 hCoV-19/England/MILK-9A9B78/2020|EPI_ISL_551038|20 ···················· ···················· ····················· ···················· ···················· ····················
J 2077 hCoV-19/England/MILK-9AAE7E/2020|EPI_ISL_551039|20 ···················· ···················· ····················· ···················· ···················· ····················
J 2078 hCoV-19/England/MILK-9AB38D/2020|EPI_ISL_551040|20 ···················· ···················· ····················· ···················· ···················· ····················
J 2079 hCoV-19/England/MILK-9A8F6E/2020|EPI_ISL_551041|20 ···················· ···················· ····················· ···················· ···················· ····················
J 2080 hCoV-19/England/MILK-9A920A/2020|EPI_ISL_551043|20 ···················· ···················· ····················· ···················· ···················· ····················
J 2081 hCoV-19/Wales/MILK-9A9D90/2020|EPI_ISL_551044|2020 ···················· ···················· ····················· ···················· ···················· ····················
J 2082 hCoV-19/England/MILK-9A94F5/2020|EPI_ISL_551045|20 ···················· ···················· ····················· ···················· ···················· ····················
J 2083 hCoV-19/England/MILK-9A9E33/2020|EPI_ISL_551046|20 ···················· ·A·················· ····················· ···················· ···················· ··A·················
J 2084 hCoV-19/England/MILK-9A9E8E/2020|EPI_ISL_551048|20 ···················· ·A·················· ····················· ···················· ···················· ··A·················
J 2085 hCoV-19/England/MILK-9A96E0/2020|EPI_ISL_551049|20 ···················· ···················· ····················· ···················· ···················· ····················
J 2086 hCoV-19/England/MILK-9A90F1/2020|EPI_ISL_551050|20 ···················· ···················· ····················· ···················· ···················· ····················
J 2087 hCoV-19/England/MILK-9A9AF3/2020|EPI_ISL_551052|20 ···················· ···················· ····················· ···················· ···················· ····················
J 2088 hCoV-19/England/MILK-9AB332/2020|EPI_ISL_551053|20 ···················· ···················· ····················· ···················· ···················· ····················
J 2089 hCoV-19/England/MILK-9AB55A/2020|EPI_ISL_551054|20 ···················· ·A·················· ····················· ···················· ···················· ····················
J 2090 hCoV-19/England/MILK-9AAE14/2020|EPI_ISL_551055|20 ···················· ···················· ····················· ···················· ···················· ····················
J 2091 hCoV-19/England/MILK-9AB3C9/2020|EPI_ISL_551056|20 ···················· ···················· ····················· ···················· ···················· ····················
J 2092 hCoV-19/Wales/MILK-9A94AA/2020|EPI_ISL_551057|2020 ···················· ···················· ····················· ···················· ···················· ····················
J 2093 hCoV-19/England/MILK-994202/2020|EPI_ISL_551700|20 ···················· ···················· ····················· ···················· ···················· ····················
J 2094 hCoV-19/England/MILK-995C8B/2020|EPI_ISL_551701|20 ···················· ···················· ····················· ···················· ···················· ····················
J 2095 hCoV-19/England/MILK-995641/2020|EPI_ISL_551702|20 ···················· ···················· ····················· ···················· ···················· ····················
J 2096 hCoV-19/England/MILK-993FA2/2020|EPI_ISL_551703|20 ···················· ···················· ····················· ···················· ···················· ····················
J 2097 hCoV-19/England/MILK-993DE4/2020|EPI_ISL_551704|20 ···················· ···················· ····················· ···················· ···················· ····················
J 2098 hCoV-19/England/MILK-99435A/2020|EPI_ISL_551705|20 ···················· ·A·················· ····················· ···················· ···················· ··A·················
J 2099 hCoV-19/England/MILK-9942A8/2020|EPI_ISL_551707|20 ···················· ···················· ····················· ···················· ···················· ····················
J 2100 hCoV-19/England/MILK-993E1E/2020|EPI_ISL_551708|20 ···················· ···················· ····················· ···················· ···················· ····················
J 2101 hCoV-19/England/MILK-9956E7/2020|EPI_ISL_551709|20 ··················T· ··········M········· ····················· ···················· ·····T·············· ···················· 1
J 2102 hCoV-19/England/MILK-995571/2020|EPI_ISL_551710|20 ···················· ···················· ····················· ···················· ···················· ····················
J 2103 hCoV-19/England/MILK-99559F/2020|EPI_ISL_551711|20 ···················· ···················· ····················· ···················· ···················· ····················
J 2104 hCoV-19/England/MILK-99430F/2020|EPI_ISL_551712|20 ···················· ···················· ····················· ···················· ···················· ····················
J 2105 hCoV-19/England/MILK-993EA5/2020|EPI_ISL_551713|20 ···················· ···················· ····················· ···················· ···················· ····················
J 2106 hCoV-19/England/MILK-9956C9/2020|EPI_ISL_551715|20 ···················· ···················· ····················· ···················· ···················· ····················
J 2107 hCoV-19/Wales/MILK-993F0C/2020|EPI_ISL_551716|2020 ···················· ···················· ····················· ···················· ···················· ····················
J 2108 hCoV-19/England/MILK-995BBB/2020|EPI_ISL_551717|20 ···················· ···················· ····················· ···················· ···················· ····················
J 2109 hCoV-19/England/MILK-9943F0/2020|EPI_ISL_551718|20 ···················· ···················· ····················· ···················· ···················· ····················
J 2110 hCoV-19/England/MILK-993BAE/2020|EPI_ISL_552300|20 ···················· ·A·················· ····················· ···················· ···················· ··A·················
J 2111 hCoV-19/England/MILK-994BDA/2020|EPI_ISL_552356|20 ···················· ·A·················· ····················· ···················· ···················· ··A·················
J 2112 hCoV-19/England/MILK-994633/2020|EPI_ISL_552357|20 ···················· ···················· ····················· ···················· ···················· ····················
J 2113 hCoV-19/Wales/MILK-99391D/2020|EPI_ISL_552358|2020 ···················· ···················· ····················· ···················· ···················· ····················
J 2114 hCoV-19/England/MILK-993BEA/2020|EPI_ISL_552361|20 ···················· ···················· ····················· ···················· ···················· ····················
J 2115 hCoV-19/England/MILK-994CF5/2020|EPI_ISL_552362|20 ···················· ···················· ····················· ···················· ···················· ····················
J 2116 hCoV-19/England/MILK-994897/2020|EPI_ISL_552363|20 ···················· ···················· ····················· ···················· ···················· ····················
J 2117 hCoV-19/England/MILK-994A91/2020|EPI_ISL_552365|20 ···················· ···················· ····················· ···················· ···················· ····················
J 2118 hCoV-19/England/MILK-9945DC/2020|EPI_ISL_552367|20 ···················· ···················· ····················· ···················· ···················· ····················
J 2119 hCoV-19/England/MILK-991D22/2020|EPI_ISL_552368|20 ···················· ···················· ····················· ···················· ···················· ····················
J 2120 hCoV-19/England/MILK-995025/2020|EPI_ISL_552369|20 ···················· ···················· ····················· ···················· ···················· ····················
J 2121 hCoV-19/England/MILK-994D5C/2020|EPI_ISL_552370|20 ···················· ···················· ····················· ···················· ···················· ····················
J 2122 hCoV-19/England/MILK-99392C/2020|EPI_ISL_552371|20 ···················· ···················· ····················· ···················· ···················· ····················
J 2123 hCoV-19/England/MILK-995492/2020|EPI_ISL_552372|20 ···················· ···················· ····················· ···················· ···················· ····················
J 2124 hCoV-19/England/MILK-9949C1/2020|EPI_ISL_552373|20 ···················· ···················· ····················· ···················· ···················· ····················
J 2125 hCoV-19/England/MILK-991E01/2020|EPI_ISL_552375|20 ···················· ···················· ····················· ···················· ···················· ····················
J 2126 hCoV-19/England/MILK-994703/2020|EPI_ISL_552376|20 ···················· ···················· ····················· ···················· ···················· ····················
J 2127 hCoV-19/England/MILK-99479A/2020|EPI_ISL_552378|20 ···················· ···················· ····················· ···················· ···················· ····················
J 2128 hCoV-19/England/MILK-994F0B/2020|EPI_ISL_552380|20 ···················· ···················· ····················· ···················· ···················· ····················
J 2129 hCoV-19/England/MILK-993A83/2020|EPI_ISL_552381|20 ···················· ···················· ····················· ···················· ···················· ····················
J 2130 hCoV-19/England/MILK-994D89/2020|EPI_ISL_552382|20 ···················· ·A·················· ····················· ···················· ···················· ··A·················
J 2131 hCoV-19/England/MILK-993D5D/2020|EPI_ISL_552383|20 ···················· ···················· ····················· ···················· ···················· ····················
J 2132 hCoV-19/England/MILK-994985/2020|EPI_ISL_552384|20 ···················· ···················· ····················· ···················· ···················· ····················
J 2133 hCoV-19/Wales/MILK-994C13/2020|EPI_ISL_552385|2020 ···················· ···················· ····················· ···················· ···················· ····················
J 2134 hCoV-19/England/MILK-994A46/2020|EPI_ISL_552386|20 ···················· ···················· ····················· ···················· ···················· ····················
J 2135 hCoV-19/England/MILK-993AB0/2020|EPI_ISL_552387|20 ···················· ···················· ····················· ···················· ···················· ····················

| | | | | | | | | |
|---|---|---|---|---|---|---|---|---|
| J | 2136 | hCoV-19/England/ALDP-955003/2020\|EPI_ISL_553600\|20 | ···················· | ·A·················· | ···················· | ···················· | ···················· | ···················· |
| J | 2137 | hCoV-19/England/ALDP-9553DD/2020\|EPI_ISL_553602\|20 | ···················· | ···················· | ···················· | ···················· | ···················· | ···················· |
| J | 2138 | hCoV-19/England/ALDP-954F07/2020\|EPI_ISL_553603\|20 | ···················· | ···················· | ···················· | ···················· | ···················· | ···················· |
| J | 2139 | hCoV-19/England/ALDP-95CBFC/2020\|EPI_ISL_553604\|20 | ···················· | ···················· | ···················· | ···················· | ···················· | ···················· |
| J | 2140 | hCoV-19/England/ALDP-9551F1/2020\|EPI_ISL_553605\|20 | ···················· | ···················· | ···················· | ···················· | ···················· | ···················· |
| J | 2141 | hCoV-19/England/ALDP-95A94F/2020\|EPI_ISL_553606\|20 | ···················· | ···················· | ···················· | ···················· | ···················· | ···················· |
| J | 2142 | hCoV-19/England/ALDP-954A06/2020\|EPI_ISL_553607\|20 | ···················· | ···················· | ···················· | ···················· | ···················· | ···················· |
| J | 2143 | hCoV-19/England/ALDP-954FE9/2020\|EPI_ISL_553608\|20 | ···················· | ···················· | ···················· | ···················· | ···················· | ···················· |
| J | 2144 | hCoV-19/England/ALDP-95D135/2020\|EPI_ISL_553609\|20 | ···················· | ···················· | ···················· | ···················· | ···················· | ···················· |
| J | 2145 | hCoV-19/England/ALDP-95CAD1/2020\|EPI_ISL_553610\|20 | ···················· | ···················· | ···················· | ···················· | ···················· | ···················· |
| J | 2146 | hCoV-19/Wales/ALDP-94D6A2/2020\|EPI_ISL_553611\|2020 | ···················· | ···················· | ···················· | ···················· | ···················· | ···················· |
| J | 2147 | hCoV-19/England/ALDP-95CA59/2020\|EPI_ISL_553612\|20 | ···················· | ·············A······ | ···················· | ···················· | ···················· | ···················· |
| J | 2148 | hCoV-19/England/ALDP-95A4D5/2020\|EPI_ISL_553613\|20 | ···················· | ···················· | ···················· | ···················· | ···················· | ···················· |
| J | 2149 | hCoV-19/England/ALDP-9555E6/2020\|EPI_ISL_553614\|20 | ···················· | ···················· | ···················· | ···················· | ···················· | ···················· |
| J | 2150 | hCoV-19/England/ALDP-955258/2020\|EPI_ISL_553615\|20 | ···················· | ···················· | ···················· | ···················· | ···················· | ···················· |
| J | 2151 | hCoV-19/England/ALDP-95C989/2020\|EPI_ISL_553616\|20 | ···················· | ···················· | ···················· | ···················· | ···················· | ···················· |
| J | 2152 | hCoV-19/Wales/ALDP-95CC71/2020\|EPI_ISL_553617\|2020 | ···················· | ···················· | ···················· | ···················· | ···················· | ···················· |
| J | 2153 | hCoV-19/England/ALDP-9553EC/2020\|EPI_ISL_553618\|20 | ···················· | ···················· | ···················· | ···················· | ···················· | ···················· |
| J | 2154 | hCoV-19/England/ALDP-959FD5/2020\|EPI_ISL_553619\|20 | ···················· | ···················· | ···················· | ···················· | ···················· | ···················· |
| J | 2155 | hCoV-19/England/ALDP-95523A/2020\|EPI_ISL_553620\|20 | ···················· | ···················· | ···················· | ···················· | ···················· | ···················· |
| J | 2156 | hCoV-19/England/ALDP-95520D/2020\|EPI_ISL_553621\|20 | ···················· | ···················· | ···················· | ···················· | ···················· | ···················· |
| J | 2157 | hCoV-19/England/ALDP-95555F/2020\|EPI_ISL_553622\|20 | ···················· | ···················· | ···················· | ···················· | ···················· | ···················· |
| J | 2158 | hCoV-19/England/ALDP-954BE5/2020\|EPI_ISL_553623\|20 | ···················· | ···················· | ···················· | ···················· | ···················· | ···················· |
| J | 2159 | hCoV-19/England/ALDP-955610/2020\|EPI_ISL_553624\|20 | ···················· | ···················· | ···················· | ···················· | ···················· | ···················· |
| J | 2160 | hCoV-19/England/ALDP-95CC08/2020\|EPI_ISL_553625\|20 | ···················· | ···················· | ···················· | ···················· | ···················· | ···················· |
| J | 2161 | hCoV-19/England/ALDP-95C9B6/2020\|EPI_ISL_553627\|20 | ···················· | ···················· | ···················· | ···················· | ···················· | ···················· |
| J | 2162 | hCoV-19/England/ALDP-9554BC/2020\|EPI_ISL_553629\|20 | ···················· | ···················· | ···················· | ···················· | ···················· | ···················· |
| J | 2163 | hCoV-19/England/ALDP-954FF8/2020\|EPI_ISL_553630\|20 | ···················· | ···················· | ···················· | ···················· | ···················· | ···················· |
| J | 2164 | hCoV-19/England/ALDP-9520AC/2020\|EPI_ISL_553631\|20 | ···················· | ···················· | ···················· | ···················· | ···················· | ···················· |
| J | 2165 | hCoV-19/England/ALDP-95225B/2020\|EPI_ISL_553687\|20 | ···················· | ···················· | ···················· | ···················· | ···················· | ···················· |
| J | 2166 | hCoV-19/England/ALDP-9521B8/2020\|EPI_ISL_553688\|20 | ···················· | ···················· | ···················· | ···················· | ···················· | ···················· |
| J | 2167 | hCoV-19/England/ALDP-95216D/2020\|EPI_ISL_553691\|20 | ···················· | ···················· | ···················· | ···················· | ···················· | ···················· |
| J | 2168 | hCoV-19/England/ALDP-9530C9/2020\|EPI_ISL_553693\|20 | ···················· | ···················· | ···················· | ···················· | ···················· | ···················· |
| J | 2169 | hCoV-19/England/ALDP-952F09/2020\|EPI_ISL_553694\|20 | ···················· | ···················· | ···················· | ···················· | ···················· | ···················· |
| J | 2170 | hCoV-19/England/ALDP-952DA5/2020\|EPI_ISL_553695\|20 | ···················· | ···················· | ···················· | ···················· | ···················· | ···················· |
| J | 2171 | hCoV-19/England/ALDP-95306F/2020\|EPI_ISL_553696\|20 | ···················· | ···················· | ···················· | ···················· | ···················· | ···················· |
| J | 2172 | hCoV-19/England/ALDP-952E84/2020\|EPI_ISL_553697\|20 | ···················· | ···················· | ···················· | ···················· | ···················· | ···················· |
| J | 2173 | hCoV-19/England/ALDP-952279/2020\|EPI_ISL_553698\|20 | ···················· | ···················· | ···················· | ···················· | ···················· | ···················· |
| J | 2174 | hCoV-19/England/ALDP-95A244/2020\|EPI_ISL_554900\|20 | ···················· | ···················· | ···················· | ···················· | ···················· | ···················· |
| J | 2175 | hCoV-19/England/ALDP-95A0B3/2020\|EPI_ISL_554901\|20 | ···················· | ···················· | ···········T········ | ···················· | ···················· | ···················· |
| J | 2176 | hCoV-19/England/ALDP-94E489/2020\|EPI_ISL_554902\|20 | ···················· | ···················· | ···················· | ···················· | ···················· | ···················· |
| J | 2177 | hCoV-19/England/ALDP-95ABB2/2020\|EPI_ISL_554903\|20 | ···················· | ···················· | ···················· | ···················· | ···················· | ···················· |
| J | 2178 | hCoV-19/England/ALDP-95C1BE/2020\|EPI_ISL_554904\|20 | ···················· | ···················· | ···················· | ···················· | ···················· | ···················· |
| J | 2179 | hCoV-19/England/ALDP-95C39A/2020\|EPI_ISL_554905\|20 | ···················· | ···················· | ···················· | ···················· | ···················· | ···················· |
| J | 2180 | hCoV-19/Wales/ALDP-95AB1C/2020\|EPI_ISL_554908\|2020 | ···················· | ···················· | ···················· | ···················· | ···················· | ···················· |
| J | 2181 | hCoV-19/England/ALDP-95CF78/2020\|EPI_ISL_554909\|20 | ···················· | ···················· | ···················· | ···················· | ···················· | ···················· |
| J | 2182 | hCoV-19/England/ALDP-95A7AF/2020\|EPI_ISL_554910\|20 | ···················· | ···················· | ···················· | ···················· | ···················· | ···················· |
| J | 2183 | hCoV-19/England/ALDP-94D5A5/2020\|EPI_ISL_554911\|20 | ···················· | ···················· | ···················· | ···················· | ···················· | ···················· |
| J | 2184 | hCoV-19/England/ALDP-95CCEA/2020\|EPI_ISL_554912\|20 | ···················· | ···················· | ···················· | ···················· | ···················· | ···················· |
| J | 2185 | hCoV-19/England/ALDP-95AC46/2020\|EPI_ISL_554913\|20 | ···················· | ···················· | ···················· | ···················· | ···················· | ···················· |
| J | 2186 | hCoV-19/England/ALDP-95AAC4/2020\|EPI_ISL_554914\|20 | ···················· | ···················· | ···················· | ···················· | ···················· | ···················· |
| J | 2187 | hCoV-19/England/ALDP-95C36D/2020\|EPI_ISL_554915\|20 | ···················· | ···················· | ···················· | ···················· | ···················· | ···················· |
| J | 2188 | hCoV-19/England/ALDP-95CFE1/2020\|EPI_ISL_554917\|20 | ···················· | ···················· | ···················· | ···················· | ···················· | ···················· |
| J | 2189 | hCoV-19/England/ALDP-95A61B/2020\|EPI_ISL_554918\|20 | ···················· | ···················· | ···················· | ···················· | ···················· | ···················· |
| J | 2190 | hCoV-19/Wales/ALDP-95AAD3/2020\|EPI_ISL_554919\|2020 | ···················· | ···················· | ···················· | ···················· | ···················· | ···················· |
| J | 2191 | hCoV-19/England/ALDP-95C38B/2020\|EPI_ISL_554920\|20 | ···················· | ···················· | ···················· | ···················· | ···················· | ···················· |
| J | 2192 | hCoV-19/England/ALDP-94D815/2020\|EPI_ISL_554921\|20 | ···················· | ···················· | ···················· | ···················· | ···················· | ···················· |
| J | 2193 | hCoV-19/England/ALDP-95C181/2020\|EPI_ISL_554922\|20 | ···················· | ·A·················· | ···················· | ···················· | ···················· | ···················· |
| J | 2194 | hCoV-19/England/ALDP-94E771/2020\|EPI_ISL_554924\|20 | ···················· | ···················· | ···················· | ···················· | ···················· | ···················· |
| J | 2195 | hCoV-19/England/ALDP-95C7AD/2020\|EPI_ISL_554925\|20 | ···················· | ···················· | ···················· | ···················· | ···················· | ···················· |
| J | 2196 | hCoV-19/England/ALDP-95C42E/2020\|EPI_ISL_554926\|20 | ···················· | ···················· | ···················· | ···················· | ···················· | ···················· |
| J | 2197 | hCoV-19/England/ALDP-95C488/2020\|EPI_ISL_554927\|20 | ···················· | ···················· | ···················· | ···················· | ···················· | ···················· |
| J | 2198 | hCoV-19/England/ALDP-959E9C/2020\|EPI_ISL_554928\|20 | ···················· | ···················· | ···················· | ···················· | ···················· | ···················· |
| J | 2199 | hCoV-19/England/ALDP-95ABC1/2020\|EPI_ISL_554931\|20 | ···················· | ···················· | ···················· | ···················· | ···················· | ···················· |
| J | 2200 | hCoV-19/England/ALDP-95C303/2020\|EPI_ISL_554932\|20 | ···················· | ···················· | ···················· | ···················· | ···················· | ···················· |
| J | 2201 | hCoV-19/England/ALDP-95C1EB/2020\|EPI_ISL_554934\|20 | ···················· | ···················· | ···················· | ···················· | ···················· | ···················· |
| J | 2202 | hCoV-19/England/ALDP-94E97B/2020\|EPI_ISL_554935\|20 | ···················· | ···················· | ···················· | ···················· | ···················· | ···················· |
| J | 2203 | hCoV-19/England/ALDP-94E911/2020\|EPI_ISL_554937\|20 | ···················· | ···················· | ···················· | ···················· | ···················· | ···················· |
| J | 2204 | hCoV-19/England/ALDP-95CC26/2020\|EPI_ISL_554938\|20 | ···················· | ···················· | ···················· | ···················· | ···················· | ···················· |
| J | 2205 | hCoV-19/England/ALDP-95A0A4/2020\|EPI_ISL_554939\|20 | ···················· | ···················· | ···················· | ···················· | ···················· | ···················· |
| J | 2206 | hCoV-19/England/ALDP-95C901/2020\|EPI_ISL_554940\|20 | ···················· | ···················· | ···················· | ···················· | ···················· | ···················· |
| J | 2207 | hCoV-19/England/ALDP-95C5EF/2020\|EPI_ISL_554941\|20 | ···················· | ·A·················· | ···················· | ···················· | ···················· | ···················· |
| J | 2208 | hCoV-19/England/ALDP-95CF69/2020\|EPI_ISL_554942\|20 | ···················· | ···················· | ···················· | ···················· | ···················· | ···················· |
| J | 2209 | hCoV-19/England/ALDP-95CE4E/2020\|EPI_ISL_554943\|20 | ···················· | ···················· | ···················· | ···················· | ···················· | ···················· |
| J | 2210 | hCoV-19/England/ALDP-94EA69/2020\|EPI_ISL_554944\|20 | ···················· | ·A·················· | ···················· | ···················· | ···················· | ···A················ |
| J | 2211 | hCoV-19/England/ALDP-95C109/2020\|EPI_ISL_554945\|20 | ···················· | ···················· | ···················· | ···················· | ···················· | ···················· |
| J | 2212 | hCoV-19/England/ALDP-95C4B5/2020\|EPI_ISL_554946\|20 | ···················· | ·A·················· | ···················· | ···················· | ···················· | ···················· |
| J | 2213 | hCoV-19/England/ALDP-95C479/2020\|EPI_ISL_554947\|20 | ···················· | ···················· | ···················· | ···················· | ···················· | ···················· |
| J | 2214 | hCoV-19/England/ALDP-95AC64/2020\|EPI_ISL_554948\|20 | ···················· | ···················· | ···················· | ···················· | ···················· | ···················· |
| J | 2215 | hCoV-19/England/ALDP-94D7DC/2020\|EPI_ISL_554950\|20 | ···················· | ···················· | ···················· | ···················· | ···················· | ···················· |

| | | | | | | | | |
|---|---|---|---|---|---|---|---|---|
| J | 2216 | hCoV-19/England/ALDP-95C637/2020\|EPI_ISL_554951\|20 | ···················· | ···················· | ····················· | ···················· | ···················· | ···················· |
| J | 2217 | hCoV-19/England/ALDP-94D657/2020\|EPI_ISL_554952\|20 | ···················· | ···················· | ····················· | ···················· | ···················· | ···················· |
| J | 2218 | hCoV-19/England/ALDP-94D851/2020\|EPI_ISL_554953\|20 | ···················· | ···················· | ····················· | ···················· | ···················· | ···················· |
| J | 2219 | hCoV-19/England/ALDP-95C43D/2020\|EPI_ISL_554955\|20 | ···················· | ···················· | ····················· | ···················· | ···················· | ···················· |
| J | 2220 | hCoV-19/England/ALDP-95C3B8/2020\|EPI_ISL_554956\|20 | ···················· | ···················· | ····················· | ···················· | ···················· | ···················· |
| J | 2221 | hCoV-19/England/ALDP-95C5B2/2020\|EPI_ISL_554960\|20 | ···················· | ···················· | ····················· | ···················· | ···················· | ···················· |
| J | 2222 | hCoV-19/England/ALDP-94E7BD/2020\|EPI_ISL_554961\|20 | ···················· | ···················· | ····················· | ···················· | ···················· | ···················· |
| J | 2223 | hCoV-19/England/ALDP-94F85F/2020\|EPI_ISL_555260\|20 | ···················· | ···················· | ····················· | ···················· | ···················· | ···················· |
| J | 2224 | hCoV-19/England/ALDP-94FF2D/2020\|EPI_ISL_555261\|20 | ···················· | ···················· | ····················· | ···················· | ···················· | ···················· |
| J | 2225 | hCoV-19/England/ALDP-94F94D/2020\|EPI_ISL_555262\|20 | ···················· | ···················· | ····················· | ···················· | ···················· | ···················· |
| J | 2226 | hCoV-19/England/ALDP-9500CC/2020\|EPI_ISL_555263\|20 | ···················· | ···················· | ····················· | ···················· | ···················· | ···················· |
| J | 2227 | hCoV-19/England/ALDP-94DA2E/2020\|EPI_ISL_555264\|20 | ···················· | ···················· | ····················· | ···················· | ···················· | ···················· |
| J | 2228 | hCoV-19/England/ALDP-94FB92/2020\|EPI_ISL_555265\|20 | ···················· | ···················· | ····················· | ···················· | ···················· | ···················· |
| J | 2229 | hCoV-19/England/ALDP-94FE3F/2020\|EPI_ISL_555267\|20 | ···················· | ···················· | ····················· | ···················· | ···················· | ···················· |
| J | 2230 | hCoV-19/England/ALDP-94E146/2020\|EPI_ISL_555268\|20 | ···················· | ···················· | ····················· | ···················· | ···················· | ···················· |
| J | 2231 | hCoV-19/England/ALDP-94E717/2020\|EPI_ISL_555269\|20 | ···················· | ·A·················· | ····················· | ···················· | ···················· | ···················· |
| J | 2232 | hCoV-19/Wales/ALDP-9503F0/2020\|EPI_ISL_555270\|2020 | ···················· | ···················· | ····················· | ···················· | ···················· | ···················· |
| J | 2233 | hCoV-19/England/ALDP-94FDE7/2020\|EPI_ISL_555271\|20 | ···················· | ···················· | ····················· | ···················· | ···················· | ···················· |
| J | 2234 | hCoV-19/England/ALDP-94F9D4/2020\|EPI_ISL_555272\|20 | ···················· | ···················· | ····················· | ···················· | ···················· | ···················· |
| J | 2235 | hCoV-19/England/ALDP-95028A/2020\|EPI_ISL_555273\|20 | ···················· | ···················· | ····················· | ···················· | ···················· | ···················· |
| J | 2236 | hCoV-19/England/ALDP-94FD7E/2020\|EPI_ISL_555275\|20 | ···················· | ···················· | ····················· | ···················· | ···················· | ···················· |
| J | 2237 | hCoV-19/England/ALDP-94D9C7/2020\|EPI_ISL_555277\|20 | ···················· | ···················· | ····················· | ···················· | ···················· | ···················· |
| J | 2238 | hCoV-19/England/ALDP-94DA97/2020\|EPI_ISL_555278\|20 | ···················· | ·A·················· | ····················· | ···················· | ···················· | ···················· |
| J | 2239 | hCoV-19/England/ALDP-94FC9F/2020\|EPI_ISL_555280\|20 | ···················· | ···················· | ····················· | ···················· | ···················· | ···················· |
| J | 2240 | hCoV-19/England/ALDP-95041B/2020\|EPI_ISL_555281\|20 | ···················· | ···················· | ····················· | ···················· | ···················· | ···················· |
| J | 2241 | hCoV-19/England/ALDP-94FC17/2020\|EPI_ISL_555282\|20 | ···················· | ···················· | ····················· | ···················· | ···················· | ···················· |
| J | 2242 | hCoV-19/England/ALDP-950509/2020\|EPI_ISL_555283\|20 | ···················· | ···················· | ····················· | ···················· | ···················· | ···················· |
| J | 2243 | hCoV-19/England/ALDP-94DA1F/2020\|EPI_ISL_555284\|20 | ···················· | ···················· | ···········T········· | ···················· | ···················· | ···················· |
| J | 2244 | hCoV-19/England/ALDP-94FC44/2020\|EPI_ISL_555285\|20 | ···················· | ···················· | ····················· | ···················· | ···················· | ···················· |
| J | 2245 | hCoV-19/England/ALDP-94F840/2020\|EPI_ISL_555286\|20 | ···················· | ···················· | ····················· | ···················· | ···················· | ···················· |
| J | 2246 | hCoV-19/England/ALDP-950026/2020\|EPI_ISL_555287\|20 | ···················· | ·A·················· | ····················· | ···················· | ···················· | ···················· |
| J | 2247 | hCoV-19/England/ALDP-94FBFC/2020\|EPI_ISL_555289\|20 | ···················· | ···················· | ····················· | ···················· | ···················· | ···················· |
| J | 2248 | hCoV-19/England/ALDP-94FB83/2020\|EPI_ISL_555290\|20 | ···················· | ···················· | ····················· | ···················· | ···················· | ···················· |
| J | 2249 | hCoV-19/England/ALDP-9502F3/2020\|EPI_ISL_555291\|20 | ···················· | ···················· | ····················· | ···················· | ···················· | ···················· |
| J | 2250 | hCoV-19/England/ALDP-9501D8/2020\|EPI_ISL_555292\|20 | ···················· | ···················· | ····················· | ···················· | ···················· | ···················· |
| J | 2251 | hCoV-19/England/ALDP-94FF69/2020\|EPI_ISL_555293\|20 | ···················· | ···················· | ····················· | ···················· | ···················· | ···················· |
| J | 2252 | hCoV-19/England/ALDP-95030F/2020\|EPI_ISL_555294\|20 | ···················· | ···················· | ····················· | ···················· | ···················· | ···················· |
| J | 2253 | hCoV-19/England/ALDP-950062/2020\|EPI_ISL_555295\|20 | ···················· | ···················· | ····················· | ···················· | ···················· | ···················· |
| J | 2254 | hCoV-19/England/ALDP-94DCEC/2020\|EPI_ISL_555296\|20 | ···················· | ···················· | ····················· | ···················· | ···················· | ···················· |
| J | 2255 | hCoV-19/England/ALDP-94FB65/2020\|EPI_ISL_555297\|20 | ···················· | ···················· | ····················· | ···················· | ···················· | ···················· |
| J | 2256 | hCoV-19/England/ALDP-94FC62/2020\|EPI_ISL_555298\|20 | ···················· | ···················· | ····················· | ···················· | ···················· | ···················· |
| J | 2257 | hCoV-19/Wales/ALDP-94FF78/2020\|EPI_ISL_555299\|2020 | ···················· | ···················· | ····················· | ···················· | ···················· | ···················· |
| J | 2258 | hCoV-19/England/ALDP-8F79EB/2020\|EPI_ISL_556591\|20 | ···················· | ···················· | ····················· | ···················· | ···················· | ···················· |
| J | 2259 | hCoV-19/England/ALDP-8CABBE/2020\|EPI_ISL_556592\|20 | ···················· | ···················· | ····················· | ···················· | ···················· | ···················· |
| J | 2260 | hCoV-19/England/ALDP-8E49F0/2020\|EPI_ISL_556594\|20 | ···················· | ···················· | ····················· | ···················· | ···················· | ···················· |
| J | 2261 | hCoV-19/England/ALDP-8E328D/2020\|EPI_ISL_556595\|20 | ···················· | ···················· | ····················· | ···················· | ···················· | ···················· |
| J | 2262 | hCoV-19/England/ALDP-8E5397/2020\|EPI_ISL_556597\|20 | ···················· | ·A·················· | ····················· | ···················· | ···················· | ···················· |
| J | 2263 | hCoV-19/England/ALDP-8C12B2/2020\|EPI_ISL_556598\|20 | ···················· | ···················· | ····················· | ···················· | ···················· | ···················· |
| J | 2264 | hCoV-19/England/ALDP-8E68B5/2020\|EPI_ISL_556599\|20 | ···················· | ···················· | ····················· | ···················· | ···················· | ···················· |
| J | 2265 | hCoV-19/England/ALDP-8D736B/2020\|EPI_ISL_557160\|20 | ···················· | ···················· | ····················· | ···················· | ···················· | ···················· |
| J | 2266 | hCoV-19/England/ALDP-90DA39/2020\|EPI_ISL_557162\|20 | ···················· | ···················· | ····················· | ···················· | ···················· | ···················· |
| J | 2267 | hCoV-19/England/ALDP-8D5619/2020\|EPI_ISL_557163\|20 | ···················· | ···················· | ····················· | ···················· | ···················· | ···················· |
| J | 2268 | hCoV-19/England/ALDP-8FFDBA/2020\|EPI_ISL_557164\|20 | ···················· | ···················· | ····················· | ···················· | ···················· | ···················· |
| J | 2269 | hCoV-19/England/ALDP-8B9E6E/2020\|EPI_ISL_557165\|20 | ···················· | ···················· | ····················· | ···················· | ···················· | ···················· |
| J | 2270 | hCoV-19/England/MILK-81BEA6/2020\|EPI_ISL_557167\|20 | ···················· | ···················· | ····················· | ···················· | ···················· | ···················· |
| J | 2271 | hCoV-19/England/MILK-878C34/2020\|EPI_ISL_557168\|20 | ···················· | ·A·················· | ····················· | ···················· | ···················· | ···················· |
| J | 2272 | hCoV-19/England/MILK-84717B/2020\|EPI_ISL_557170\|20 | ···················· | ···················· | ····················· | ···················· | ···················· | ···················· |
| J | 2273 | hCoV-19/England/MILK-80EFB2/2020\|EPI_ISL_557171\|20 | ···················· | ···················· | ····················· | ···················· | ···················· | ···················· |
| J | 2274 | hCoV-19/England/MILK-803C6B/2020\|EPI_ISL_557172\|20 | ···················· | ···················· | ····················· | ···················· | ···················· | ···················· |
| J | 2275 | hCoV-19/England/MILK-87A300/2020\|EPI_ISL_557173\|20 | ···················· | ·A·················· | ····················· | ···················· | ···················· | ···················· |
| J | 2276 | hCoV-19/England/MILK-8305A6/2020\|EPI_ISL_557174\|20 | ···················· | ···················· | ····················· | ···················· | ···················· | ···················· |
| J | 2277 | hCoV-19/England/MILK-83247A/2020\|EPI_ISL_557175\|20 | ···················· | ···················· | ····················· | ···················· | ···················· | ···················· |
| J | 2278 | hCoV-19/England/MILK-840B47/2020\|EPI_ISL_557177\|20 | ···················· | ·A·················· | ····················· | ···················· | ···················· | ···················· |
| J | 2279 | hCoV-19/England/MILK-80D1BD/2020\|EPI_ISL_557178\|20 | ···················· | ···················· | ····················· | ···················· | ···················· | ···················· |
| J | 2280 | hCoV-19/England/MILK-83A964/2020\|EPI_ISL_557179\|20 | ···················· | ···················· | ····················· | ···················· | ···················· | ···················· |
| J | 2281 | hCoV-19/England/MILK-87B008/2020\|EPI_ISL_557181\|20 | ···················· | ···················· | ····················· | ···················· | ···················· | ···················· |
| J | 2282 | hCoV-19/England/MILK-7FF920/2020\|EPI_ISL_557182\|20 | ···················· | ···················· | ····················· | ···················· | ···················· | ···················· |
| J | 2283 | hCoV-19/England/MILK-837C9B/2020\|EPI_ISL_557183\|20 | ···················· | ···················· | ····················· | ···················· | ···················· | ···················· |
| J | 2284 | hCoV-19/England/MILK-84D755/2020\|EPI_ISL_557184\|20 | ···················· | ···················· | ····················· | ···················· | ···················· | ···················· |
| J | 2285 | hCoV-19/England/MILK-800527/2020\|EPI_ISL_557185\|20 | ···················· | ···················· | ····················· | ···················· | ···················· | ···················· |
| J | 2286 | hCoV-19/England/MILK-872CEF/2020\|EPI_ISL_557187\|20 | ···················· | ···················· | ····················· | ···················· | ···················· | ···················· |
| J | 2287 | hCoV-19/England/MILK-80A675/2020\|EPI_ISL_557188\|20 | ···················· | ·········A·········· | ····················· | ···················· | ···················· | ···················· |
| J | 2288 | hCoV-19/England/MILK-818E9A/2020\|EPI_ISL_557190\|20 | ···················· | ···················· | ····················· | ···················· | ···················· | ···················· |
| J | 2289 | hCoV-19/England/MILK-848D67/2020\|EPI_ISL_557191\|20 | ···················· | ·A·················· | ····················· | ···················· | ···················· | ···················· |
| J | 2290 | hCoV-19/England/MILK-81EA81/2020\|EPI_ISL_557194\|20 | ···················· | ······A············· | ····················· | ···················· | ···················· | ···················· |
| J | 2291 | hCoV-19/England/MILK-836B71/2020\|EPI_ISL_557195\|20 | ···················· | ···················· | ····················· | ···················· | ···················· | ···················· |
| J | 2292 | hCoV-19/England/MILK-8361E8/2020\|EPI_ISL_557196\|20 | ···················· | ···················· | ····················· | ···················· | ···················· | ···················· |
| J | 2293 | hCoV-19/England/MILK-859ED9/2020\|EPI_ISL_557197\|20 | ···················· | ···················· | ····················· | ···················· | ···················· | ···················· |
| J | 2294 | hCoV-19/England/MILK-825C54/2020\|EPI_ISL_557198\|20 | ···················· | ···················· | ····················· | ···················· | ···················· | ···················· |
| J | 2295 | hCoV-19/England/ALDP-5C6D37/2020\|EPI_ISL_558490\|20 | ···················· | ···················· | ····················· | ···················· | ···················· | ···················· |

J 2296 hCoV-19/England/ALDP-5C936F/2020|EPI_ISL_558491|20 ···················· ···················· ···················· ···················· ···················· ····················
J 2297 hCoV-19/England/ALDP-5D69D6/2020|EPI_ISL_558492|20 ···················· ·············A······ ···················· ···················· ···················· ····················
J 2298 hCoV-19/England/ALDP-5D1838/2020|EPI_ISL_558493|20 ···················· ···················· ···················· ···················· ···················· ····················
J 2299 hCoV-19/England/ALDP-5C5837/2020|EPI_ISL_558494|20 ···················· ···················· ···················· ···················· ···················· ····················
J 2300 hCoV-19/England/ALDP-5CF150/2020|EPI_ISL_558495|20 ···················· ···················· ···················· ···················· ···················· ····················
J 2301 hCoV-19/England/ALDP-5D173B/2020|EPI_ISL_558496|20 ···················· ···················· ···················· ···················· ···················· ····················
J 2302 hCoV-19/England/ALDP-5C97DC/2020|EPI_ISL_558497|20 ···················· ···················· ···················· ···················· ···················· ····················
J 2303 hCoV-19/England/ALDP-5D49C9/2020|EPI_ISL_558498|20 ···················· ···················· ···················· ···················· ···················· ····················
J 2304 hCoV-19/England/ALDP-5D4738/2020|EPI_ISL_558499|20 ···················· ···················· ···················· ···················· ···················· ····················
J 2305 hCoV-19/England/CAMC-9B2365/2020|EPI_ISL_549408|20 ···················· ·A·················· ···················· ···················· ···················· ····················
J 2306 hCoV-19/England/CAMC-9B2444/2020|EPI_ISL_549409|20 ···················· ···················· ···················· ···················· ···················· ····················
J 2307 hCoV-19/England/MILK-9A871B/2020|EPI_ISL_549415|20 ···················· ···················· ···················· ···················· ···················· ····················
J 2308 hCoV-19/England/MILK-9A8739/2020|EPI_ISL_549416|20 ···················· ·A·······A·········· ···················· ···················· ···················· ····················
J 2309 hCoV-19/England/MILK-9A8784/2020|EPI_ISL_549419|20 ···················· ·A·················· ···················· ···················· ···················· ····················
J 2310 hCoV-19/England/MILK-9A8793/2020|EPI_ISL_549420|20 ···················· ···················· ···················· ···················· ···················· ····················
J 2311 hCoV-19/England/MILK-9A87A2/2020|EPI_ISL_549421|20 ···················· ·A·······A·········· ···················· ···················· ···················· ····················
J 2312 hCoV-19/England/MILK-9A87C0/2020|EPI_ISL_549422|20 ···················· ···················· ···················· ···················· ···················· ····················
J 2313 hCoV-19/England/MILK-9A87DF/2020|EPI_ISL_549423|20 ···················· ···················· ···················· ···················· ···················· ····················
J 2314 hCoV-19/England/MILK-9A87EE/2020|EPI_ISL_549424|20 ···················· ·A·······A·········· ···················· ···················· ···················· ····················
J 2315 hCoV-19/England/MILK-9A87FD/2020|EPI_ISL_549425|20 ···················· ···················· ···················· ···················· ···················· ····················
J 2316 hCoV-19/England/MILK-9A8809/2020|EPI_ISL_549426|20 ···················· ·A·······A·········· ···················· ···················· ···················· ····················
J 2317 hCoV-19/England/MILK-9A8818/2020|EPI_ISL_549427|20 ···················· ···················· ···················· ···················· ···················· ····················
J 2318 hCoV-19/England/MILK-9A8845/2020|EPI_ISL_549428|20 ···················· ···················· ···················· ···················· ···················· ····················
J 2319 hCoV-19/England/MILK-9A8854/2020|EPI_ISL_549429|20 ···················· ···················· ···················· ···················· ···················· ····················
J 2320 hCoV-19/England/MILK-9A88DC/2020|EPI_ISL_549430|20 ···················· ·A·················· ···················· ···················· ···················· ····················
J 2321 hCoV-19/England/MILK-9A88EB/2020|EPI_ISL_549431|20 ···················· ···················· ···················· ···················· ···················· ····················
J 2322 hCoV-19/England/MILK-9A8906/2020|EPI_ISL_549432|20 ···················· ···················· ···················· ···················· ···················· ····················
J 2323 hCoV-19/England/MILK-9A8915/2020|EPI_ISL_549433|20 ···················· ···················· ···················· ···················· ···················· ····················
J 2324 hCoV-19/England/MILK-9A8933/2020|EPI_ISL_549434|20 ···················· ···················· ···················· ···················· ···················· ····················
J 2325 hCoV-19/England/MILK-9A8942/2020|EPI_ISL_549435|20 ···················· ·A·······A·········· ···················· ···················· ···················· ····················
J 2326 hCoV-19/England/MILK-9A8951/2020|EPI_ISL_549436|20 ···················· ···················· ···················· ···················· ···················· ····················
J 2327 hCoV-19/England/MILK-9A8A12/2020|EPI_ISL_549439|20 ···················· ···················· ···················· ···················· ···················· ····················
J 2328 hCoV-19/Wales/MILK-99A679/2020|EPI_ISL_550400|2020 ···················· ···················· ···················· ···················· ···················· ····················
J 2329 hCoV-19/England/MILK-99B414/2020|EPI_ISL_550401|20 ···················· ···················· ···················· ···················· ···················· ····················
J 2330 hCoV-19/England/MILK-99B4F6/2020|EPI_ISL_550402|20 ···················· ···················· ···················· ···················· ···················· ····················
J 2331 hCoV-19/England/MILK-99A3CD/2020|EPI_ISL_550403|20 ···················· ···················· ···················· ···················· ···················· ····················
J 2332 hCoV-19/England/MILK-99A3BE/2020|EPI_ISL_550404|20 ···················· ···················· ···················· ···················· ···················· ····················
J 2333 hCoV-19/England/MILK-99B942/2020|EPI_ISL_550405|20 ···················· ···················· ···················· ···················· ···················· ····················
J 2334 hCoV-19/England/MILK-99A688/2020|EPI_ISL_550406|20 ···················· ···················· ···················· ···················· ···················· ····················
J 2335 hCoV-19/England/MILK-99A354/2020|EPI_ISL_550407|20 ···················· ···················· ···················· ···················· ···················· ····················
J 2336 hCoV-19/England/MILK-99A57C/2020|EPI_ISL_550408|20 ···················· ···················· ···················· ···················· ···················· ····················
J 2337 hCoV-19/England/MILK-99A8CE/2020|EPI_ISL_550409|20 ···················· ···················· ···················· ···················· ···················· ····················
J 2338 hCoV-19/England/MILK-978705/2020|EPI_ISL_550486|20 ···················· ···················· ···················· ···················· ···················· ····················
J 2339 hCoV-19/England/MILK-977D40/2020|EPI_ISL_550487|20 ···················· ···················· ···················· ···················· ···················· ····················
J 2340 hCoV-19/England/MILK-977399/2020|EPI_ISL_550488|20 ···················· ···················· ···················· ···················· ···················· ····················
J 2341 hCoV-19/England/MILK-97738A/2020|EPI_ISL_550490|20 ···················· ···················· ···················· ···················· ···················· ····················
J 2342 hCoV-19/England/MILK-978626/2020|EPI_ISL_550491|20 ···················· ···················· ···················· ···················· ···················· ····················
J 2343 hCoV-19/England/MILK-978073/2020|EPI_ISL_550492|20 ···················· ···················· ···················· ···················· ···················· ····················
J 2344 hCoV-19/England/MILK-977CE9/2020|EPI_ISL_550496|20 ···················· ·············A······ ···················· ···················· ···················· ····················
J 2345 hCoV-19/England/MILK-978A66/2020|EPI_ISL_550498|20 ···················· ···················· ···················· ···················· ···················· ····················
J 2346 hCoV-19/England/MILK-995A27/2020|EPI_ISL_551720|20 ···················· ···················· ···················· ···················· ···················· ····················
J 2347 hCoV-19/England/MILK-995711/2020|EPI_ISL_551721|20 ···················· ···················· ···················· ···················· ···················· ····················
J 2348 hCoV-19/England/MILK-995C30/2020|EPI_ISL_551722|20 ···················· ·A·················· ···················· ···················· ···················· ····················
J 2349 hCoV-19/England/MILK-9940AE/2020|EPI_ISL_551723|20 ········T··········· ···················· ···················· ···················· ···················· ····················
J 2350 hCoV-19/England/MILK-9955CC/2020|EPI_ISL_551724|20 ···················· ···················· ···················· ···················· ···················· ····················
J 2351 hCoV-19/England/MILK-993F84/2020|EPI_ISL_551726|20 ···················· ···················· ···················· ···················· ···················· ····················
J 2352 hCoV-19/England/MILK-994299/2020|EPI_ISL_551728|20 ···················· ···················· ···················· ···················· ···················· ····················
J 2353 hCoV-19/England/MILK-994123/2020|EPI_ISL_551729|20 ···················· ···················· ···················· ···················· ···················· ····················
J 2354 hCoV-19/England/MILK-9941F6/2020|EPI_ISL_551730|20 ···················· ···················· ···················· ···················· ···················· ····················
J 2355 hCoV-19/England/MILK-995B9D/2020|EPI_ISL_551731|20 ···················· ·A·················· ···················· ···················· ···················· ···A················
J 2356 hCoV-19/England/MILK-995799/2020|EPI_ISL_551732|20 ········T··········· ···················· ···················· ···················· ···················· ····················
J 2357 hCoV-19/England/MILK-9942E4/2020|EPI_ISL_551733|20 ··················T· ···················· ···················· ···················· ······T············· ···················· 1
J 2358 hCoV-19/England/MILK-9955EA/2020|EPI_ISL_551734|20 ···················· ···················· ···················· ···················· ···················· ····················
J 2359 hCoV-19/England/ALDP-680BC5/2020|EPI_ISL_551735|20 ···················· ···················· ···················· ···················· ···················· ····················
J 2360 hCoV-19/England/ALDP-680196/2020|EPI_ISL_551736|20 ···················· ···················· ···················· ···················· ···················· ····················
J 2361 hCoV-19/England/MILK-6A2B81/2020|EPI_ISL_551737|20 ···················· ···················· ···················· ···················· ···················· ····················
J 2362 hCoV-19/England/MILK-698FA0/2020|EPI_ISL_551738|20 ···················· ···················· ···················· ···················· ···················· ····················
J 2363 hCoV-19/England/MILK-6AA1B4/2020|EPI_ISL_551739|20 ···················· ···················· ···················· ···················· ···················· ····················
J 2364 hCoV-19/England/MILK-6A2CF7/2020|EPI_ISL_551750|20 ···················· ···················· ···················· ···················· ···················· ····················
J 2365 hCoV-19/England/ALDP-95031E/2020|EPI_ISL_555201|20 ···················· ·A·················· ···················· ···················· ···················· ····················
J 2366 hCoV-19/England/ALDP-94FC35/2020|EPI_ISL_555202|20 ···················· ···················· ···················· ···················· ···················· ····················
J 2367 hCoV-19/England/ALDP-94DAA6/2020|EPI_ISL_555204|20 ···················· ···················· ···················· ···················· ···················· ····················
J 2368 hCoV-19/England/ALDP-950642/2020|EPI_ISL_555207|20 ···················· ···················· ···················· ···················· ···················· ····················
J 2369 hCoV-19/England/ALDP-94D8CA/2020|EPI_ISL_555208|20 ···················· ···················· ···················· ···················· ···················· ····················
J 2370 hCoV-19/England/ALDP-9502B7/2020|EPI_ISL_555209|20 ···················· ···················· ···················· ···················· ···················· ····················
J 2371 hCoV-19/England/ALDP-94DA6A/2020|EPI_ISL_555210|20 ···················· ···················· ···················· ···················· ···················· ····················
J 2372 hCoV-19/England/ALDP-950369/2020|EPI_ISL_555211|20 ···················· ···················· ···················· ···················· ···················· ····················
J 2373 hCoV-19/England/ALDP-94FE5D/2020|EPI_ISL_555212|20 ···················· ···················· ···················· ···················· ···················· ····················
J 2374 hCoV-19/England/ALDP-94DA5B/2020|EPI_ISL_555213|20 ···················· ···················· ···················· ···················· ···················· ····················
J 2375 hCoV-19/England/ALDP-950554/2020|EPI_ISL_555214|20 ···················· ···················· ···················· ···················· ···················· ····················

J 2376 hCoV-19/England/ALDP-9501F6/2020|EPI_ISL_555215|20 ···················· ···················· ····················· ···················· ···················· ····················
J 2377 hCoV-19/England/ALDP-94DEF5/2020|EPI_ISL_555216|20 ···················· ···················· ····················· ···················· ···················· ····················
J 2378 hCoV-19/England/ALDP-94FFA5/2020|EPI_ISL_555217|20 ···················· ····A··············· ····················· ···················· ···················· ····················
J 2379 hCoV-19/England/ALDP-9502D5/2020|EPI_ISL_555218|20 ···················· ···················· ····················· ···················· ···················· ····················
J 2380 hCoV-19/England/ALDP-94F910/2020|EPI_ISL_555222|20 ···················· ···················· ····················· ···················· ···················· ····················
J 2381 hCoV-19/England/ALDP-9506E8/2020|EPI_ISL_555223|20 ···················· ···················· ····················· ···················· ···················· ····················
J 2382 hCoV-19/England/ALDP-94FCBD/2020|EPI_ISL_555224|20 ···················· ···················· ····················· ···················· ···················· ····················
J 2383 hCoV-19/England/ALDP-950703/2020|EPI_ISL_555225|20 ···················· ···················· ····················· ···················· ···················· ····················
J 2384 hCoV-19/England/ALDP-94F9B6/2020|EPI_ISL_555226|20 ···················· ·A·················· ····················· ···················· ···················· ····················
J 2385 hCoV-19/England/ALDP-94FAFF/2020|EPI_ISL_555227|20 ···················· ···················· ····················· ···················· ···················· ····················
J 2386 hCoV-19/England/ALDP-95033C/2020|EPI_ISL_555228|20 ···················· ···················· ····················· ···················· ···················· ····················
J 2387 hCoV-19/England/ALDP-94FA86/2020|EPI_ISL_555229|20 ···················· ···················· ····················· ···················· ···················· ····················
J 2388 hCoV-19/England/ALDP-94FB74/2020|EPI_ISL_555230|20 ···················· ·A·················· ····················· ···················· ···················· ····················
J 2389 hCoV-19/England/ALDP-94DAB5/2020|EPI_ISL_555231|20 ···················· ···················· ····················· ···················· ···················· ····················
J 2390 hCoV-19/England/ALDP-94DF2F/2020|EPI_ISL_555232|20 ···················· ···················· ····················· ···················· ···················· ····················
J 2391 hCoV-19/England/ALDP-9505DC/2020|EPI_ISL_555233|20 ···················· ···················· ····················· ···················· ···················· ····················
J 2392 hCoV-19/England/ALDP-94FE6C/2020|EPI_ISL_555234|20 ···················· ···················· ····················· ···················· ···················· ····················
J 2393 hCoV-19/England/ALDP-950114/2020|EPI_ISL_555235|20 ···················· ···················· ····················· ···················· ···················· ····················
J 2394 hCoV-19/Wales/ALDP-94FFC3/2020|EPI_ISL_555236|2020 ···················· ···················· ····················· ···················· ···················· ····················
J 2395 hCoV-19/England/ALDP-9503E1/2020|EPI_ISL_555238|20 ···················· ···················· ····················· ···················· ···················· ····················
J 2396 hCoV-19/England/ALDP-94FD8D/2020|EPI_ISL_555239|20 ···················· ···················· ····················· ···················· ···················· ····················
J 2397 hCoV-19/England/ALDP-95069D/2020|EPI_ISL_555240|20 ···················· ···················· ····················· ···················· ···················· ····················
J 2398 hCoV-19/England/ALDP-94FCDB/2020|EPI_ISL_555241|20 ···················· ···················· ····················· ···················· ···················· ····················
J 2399 hCoV-19/England/ALDP-94FD32/2020|EPI_ISL_555242|20 ···················· ···················· ····················· ···················· ···················· ····················
J 2400 hCoV-19/England/ALDP-950378/2020|EPI_ISL_555244|20 ···················· ·A·················· ····················· ···················· ···················· ····················
J 2401 hCoV-19/England/ALDP-94FB47/2020|EPI_ISL_555245|20 ···················· ···················· ····················· ···················· ···················· ····················
J 2402 hCoV-19/England/ALDP-94FBB0/2020|EPI_ISL_555246|20 ···················· ···················· ····················· ···················· ···················· ····················
J 2403 hCoV-19/England/ALDP-94E6FC/2020|EPI_ISL_555251|20 ···················· ···················· ····················· ···················· ···················· ····················
J 2404 hCoV-19/England/ALDP-95032D/2020|EPI_ISL_555252|20 ···················· ···················· ····················· ···················· ···················· ····················
J 2405 hCoV-19/England/ALDP-94DAC4/2020|EPI_ISL_555253|20 ···················· ···················· ····················· ···················· ···················· ····················
J 2406 hCoV-19/England/ALDP-94F95C/2020|EPI_ISL_555254|20 ···················· ···················· ····················· ···················· ···················· ····················
J 2407 hCoV-19/England/ALDP-94DF10/2020|EPI_ISL_555255|20 ···················· ···················· ····················· ···················· ···················· ····················
J 2408 hCoV-19/England/ALDP-9504EE/2020|EPI_ISL_555256|20 ···················· ···················· ····················· ···················· ···················· ····················
J 2409 hCoV-19/England/ALDP-94FA1D/2020|EPI_ISL_555257|20 ···················· ···················· ····················· ···················· ···················· ····················
J 2410 hCoV-19/England/ALDP-94FC80/2020|EPI_ISL_555258|20 ···················· ···················· ····················· ···················· ···················· ····················
J 2411 hCoV-19/England/ALDP-949882/2020|EPI_ISL_556503|20 ···················· ···················· ····················· ···················· ···················· ····················
J 2412 hCoV-19/England/ALDP-9498BF/2020|EPI_ISL_556504|20 ···················· ···················· ····················· ···················· ···················· ····················
J 2413 hCoV-19/England/ALDP-94EE12/2020|EPI_ISL_556506|20 ···················· ···················· ····················· ···················· ···················· ····················
J 2414 hCoV-19/England/ALDP-94ABF1/2020|EPI_ISL_556507|20 ···················· ···················· ····················· ···················· ···················· ····················
J 2415 hCoV-19/England/CAMC-94A326/2020|EPI_ISL_556508|20 ···················· ···················· ····················· ···················· ···················· ····················
J 2416 hCoV-19/England/CAMC-94AD37/2020|EPI_ISL_556509|20 ···················· ···················· ····················· ···················· ···················· ····················
J 2417 hCoV-19/England/ALDP-94B1B2/2020|EPI_ISL_556513|20 ···················· ···················· ····················· ···················· ···················· ····················
J 2418 hCoV-19/England/ALDP-94D068/2020|EPI_ISL_556514|20 ···················· ···················· ····················· ···················· ···················· ····················
J 2419 hCoV-19/England/ALDP-94AF31/2020|EPI_ISL_556515|20 ···················· ···················· ····················· ···················· ···················· ····················
J 2420 hCoV-19/England/CAMC-949EBD/2020|EPI_ISL_556518|20 ···················· ···················· ····················· ···················· ···················· ····················
J 2421 hCoV-19/England/CAMC-9475E7/2020|EPI_ISL_556520|20 ···················· ···················· ····················· ···················· ···················· ····················
J 2422 hCoV-19/England/CAMC-946F71/2020|EPI_ISL_556521|20 ···················· ···················· ····················· ···················· ···················· ····················
J 2423 hCoV-19/England/CAMC-948E36/2020|EPI_ISL_556522|20 ···················· ···················· ····················· ···················· ···················· ····················
J 2424 hCoV-19/England/CAMC-947A60/2020|EPI_ISL_556523|20 ···················· ···················· ····················· ···················· ···················· ····················
J 2425 hCoV-19/England/CAMC-948829/2020|EPI_ISL_556524|20 ···················· ···················· ····················· ···················· ···················· ····················
J 2426 hCoV-19/England/CAMC-9479AF/2020|EPI_ISL_556526|20 ···················· ···················· ····················· ···················· ···················· ····················
J 2427 hCoV-19/England/CAMC-948DC0/2020|EPI_ISL_556528|20 ···················· ·A·················· ····················· ···················· ···················· ····················
J 2428 hCoV-19/England/CAMC-947D67/2020|EPI_ISL_556529|20 ···················· ···················· ···········T········· ···················· ···················· ····················
J 2429 hCoV-19/England/CAMC-946F35/2020|EPI_ISL_556530|20 ···················· ···················· ····················· ···················· ···················· ····················
J 2430 hCoV-19/England/CAMC-94AC49/2020|EPI_ISL_556531|20 ···················· ···················· ····················· ···················· ···················· ····················
J 2431 hCoV-19/England/CAMC-947471/2020|EPI_ISL_556532|20 ···················· ···················· ····················· ···················· ···················· ····················
J 2432 hCoV-19/England/CAMC-947A7F/2020|EPI_ISL_556533|20 ···················· ·A·················· ····················· ···················· ···················· ···A················
J 2433 hCoV-19/England/CAMC-947AF7/2020|EPI_ISL_556534|20 ···················· ···················· ····················· ···················· ···················· ····················
J 2434 hCoV-19/England/CAMC-949372/2020|EPI_ISL_556535|20 ···················· ···················· ····················· ···················· ···················· ····················
J 2435 hCoV-19/England/CAMC-9491D2/2020|EPI_ISL_556537|20 ···················· ···················· ····················· ···················· ···················· ····················
J 2436 hCoV-19/England/CAMC-94759C/2020|EPI_ISL_556538|20 ···················· ···················· ····················· ···················· ···················· ····················
J 2437 hCoV-19/England/CAMC-94822B/2020|EPI_ISL_556542|20 ···················· ···················· ····················· ···················· ···················· ····················
J 2438 hCoV-19/England/CAMC-947EEC/2020|EPI_ISL_556543|20 ···················· ·A·················· ····················· ···················· ···················· ···A················
J 2439 hCoV-19/England/CAMC-9490B7/2020|EPI_ISL_556546|20 ···················· ···················· ····················· ···················· ···················· ····················
J 2440 hCoV-19/England/CAMC-9472D1/2020|EPI_ISL_556548|20 ···················· ···················· ····················· ···················· ···················· ····················
J 2441 hCoV-19/England/CAMC-94870E/2020|EPI_ISL_556549|20 ···················· ···················· ····················· ···················· ···················· ····················
J 2442 hCoV-19/England/CAMC-94774B/2020|EPI_ISL_556551|20 ···················· ···················· ····················· ···················· ···················· ····················
J 2443 hCoV-19/England/CAMC-9470D7/2020|EPI_ISL_556552|20 ···················· ···················· ····················· ···················· ···················· ····················
J 2444 hCoV-19/England/CAMC-947875/2020|EPI_ISL_556553|20 ···················· ···················· ····················· ···················· ···················· ····················
J 2445 hCoV-19/England/CAMC-949E62/2020|EPI_ISL_556554|20 ···················· ···················· ····················· ···················· ···················· ····················
J 2446 hCoV-19/England/CAMC-94763F/2020|EPI_ISL_556560|20 ···················· ···················· ····················· ···················· ···················· ····················
J 2448 hCoV-19/England/CAMC-949390/2020|EPI_ISL_556563|20 ···················· ···················· ····················· ···················· ···················· ····················
J 2449 hCoV-19/England/CAMC-946FDB/2020|EPI_ISL_556564|20 ···················· ···················· ····················· ···················· ···················· ····················
J 2450 hCoV-19/England/CAMC-948F33/2020|EPI_ISL_556565|20 ···················· ···················· ····················· ···················· ···················· ····················
J 2451 hCoV-19/England/CAMC-94867A/2020|EPI_ISL_556566|20 ···················· ···················· ····················· ···················· ···················· ····················
J 2452 hCoV-19/England/CAMC-949E08/2020|EPI_ISL_556567|20 ···················· ···················· ····················· ···················· ···················· ····················
J 2453 hCoV-19/England/CAMC-946FEA/2020|EPI_ISL_556568|20 ···················· ···················· ····················· ···················· ···················· ····················
J 2454 hCoV-19/England/CAMC-9473DE/2020|EPI_ISL_556569|20 ···················· ···················· ····················· ···················· ···················· ····················
J 2455 hCoV-19/England/CAMC-947004/2020|EPI_ISL_556571|20 ···················· ···················· ····················· ···················· ···················· ····················
J 2456 hCoV-19/England/CAMC-946F17/2020|EPI_ISL_556572|20 ···················· ···················· ····················· ···················· ···················· ····················

J 2457 hCoV-19/England/CAMC-9490E4/2020|EPI_ISL_556575|20 ···················· ···················· ····················· ···················· ···················· ····················
J 2458 hCoV-19/England/ALDP-8C9232/2020|EPI_ISL_556577|20 ···················· ···················· ····················· ···················· ···················· ····················
J 2459 hCoV-19/England/ALDP-8F766C/2020|EPI_ISL_556580|20 ···················· ···················· ····················· ···················· ···················· ····················
J 2460 hCoV-19/England/ALDP-8EB522/2020|EPI_ISL_556583|20 ···················· ···················· ····················· ···················· ···················· ····················
J 2461 hCoV-19/England/ALDP-8BD240/2020|EPI_ISL_556588|20 ···················· ···················· ····················· ···················· ···················· ····················
J 2462 hCoV-19/England/ALDP-8F6D95/2020|EPI_ISL_556600|20 ···················· ···················· ····················· ···················· ···················· ····················
J 2463 hCoV-19/England/ALDP-8F7602/2020|EPI_ISL_556602|20 ···················· ···················· ····················· ···················· ···················· ····················
J 2464 hCoV-19/England/ALDP-8E63A5/2020|EPI_ISL_556604|20 ···················· ···················· ····················· ···················· ···················· ····················
J 2465 hCoV-19/England/ALDP-8F6928/2020|EPI_ISL_556605|20 ···················· ···················· ····················· ···················· ···················· ····················
J 2466 hCoV-19/England/ALDP-8E617E/2020|EPI_ISL_556609|20 ···················· ···················· ····················· ···················· ···················· ····················
J 2467 hCoV-19/England/ALDP-8C8E8A/2020|EPI_ISL_556610|20 ···················· ···················· ····················· ···················· ···················· ····················
J 2468 hCoV-19/England/ALDP-8F7918/2020|EPI_ISL_556611|20 ···················· ···················· ····················· ···················· ···················· ····················
J 2469 hCoV-19/Wales/ALDP-90B0A2/2020|EPI_ISL_556613|2020 ···················· ···················· ····················· ···················· ···················· ····················
J 2470 hCoV-19/England/ALDP-8E623F/2020|EPI_ISL_556614|20 ···················· ···················· ····················· ···················· ···················· ····················
J 2471 hCoV-19/England/ALDP-8C95B1/2020|EPI_ISL_556619|20 ···················· ···················· ····················· ···················· ···················· ····················
J 2472 hCoV-19/England/ALDP-8F1CBB/2020|EPI_ISL_556620|20 ···················· ···················· ····················· ···················· ···················· ····················
J 2473 hCoV-19/England/ALDP-91D720/2020|EPI_ISL_556623|20 ···················· ···················· ····················· ···················· ···················· ····················
J 2474 hCoV-19/England/ALDP-6BD479/2020|EPI_ISL_557834|20 ···················· ···················· ····················· ···················· ···················· ····················
J 2475 hCoV-19/England/ALDP-6BAB59/2020|EPI_ISL_557835|20 ···················· ···················· ····················· ···················· ···················· ····················
J 2476 hCoV-19/England/ALDP-6C0DF0/2020|EPI_ISL_557836|20 ···················· ···················· ····················· ···················· ···················· ····················
J 2477 hCoV-19/England/ALDP-6C1B31/2020|EPI_ISL_557837|20 ···················· ···················· ····················· ···················· ···················· ····················
J 2478 hCoV-19/England/MILK-615A22/2020|EPI_ISL_557838|20 ···················· ···················· ····················· ···················· ···················· ····················
J 2479 hCoV-19/England/ALDP-6B2DD3/2020|EPI_ISL_557839|20 ···················· ···················· ····················· ···················· ···················· ····················
J 2480 hCoV-19/England/ALDP-6BE812/2020|EPI_ISL_557841|20 ···················· ···················· ····················· ···················· ···················· ····················
J 2481 hCoV-19/England/ALDP-6AEC91/2020|EPI_ISL_557842|20 ···················· ···················· ····················· ···················· ···················· ····················
J 2482 hCoV-19/England/ALDP-6BCD8E/2020|EPI_ISL_557844|20 ···················· ···················· ···········T········· ···················· ···················· ····················
J 2483 hCoV-19/England/MILK-6239BB/2020|EPI_ISL_557845|20 ···················· ···················· ····················· ···················· ···················· ····················
J 2484 hCoV-19/Wales/ALDP-6AE860/2020|EPI_ISL_557846|2020 ···················· ···················· ····················· ···················· ···················· ····················
J 2485 hCoV-19/England/MILK-61A3F5/2020|EPI_ISL_557847|20 ···················· ·A·················· ····················· ···················· ···················· ····················
J 2486 hCoV-19/England/MILK-6118B3/2020|EPI_ISL_557848|20 ···················· ···················· ····················· ···················· ···················· ····················
J 2487 hCoV-19/England/MILK-62CFB0/2020|EPI_ISL_557849|20 ···················· ···················· ····················· ···················· ···················· ····················
J 2488 hCoV-19/England/ALDP-6BF6F9/2020|EPI_ISL_557850|20 ···················· ···················· ····················· ···················· ···················· ····················
J 2489 hCoV-19/England/ALDP-6B7284/2020|EPI_ISL_557851|20 ···················· ···················· ····················· ···················· ···················· ····················
J 2490 hCoV-19/England/MILK-620866/2020|EPI_ISL_557853|20 ···················· ···················· ····················· ···················· ···················· ····················
J 2491 hCoV-19/England/MILK-62CDD4/2020|EPI_ISL_557854|20 ···················· ···················· ····················· ···················· ···················· ····················
J 2492 hCoV-19/England/ALDP-6AF261/2020|EPI_ISL_557855|20 ···················· ···················· ····················· ···················· ···················· ····················
J 2493 hCoV-19/England/ALDP-6C1CD4/2020|EPI_ISL_557856|20 ···················· ···················· ····················· ···················· ···················· ····················
J 2494 hCoV-19/England/ALDP-6AEE31/2020|EPI_ISL_557857|20 ···················· ···················· ····················· ···················· ···················· ····················
J 2495 hCoV-19/England/MILK-62DF46/2020|EPI_ISL_557858|20 ···················· ···················· ····················· ···················· ···················· ····················
J 2496 hCoV-19/England/ALDP-6C3786/2020|EPI_ISL_557860|20 ···················· ···················· ····················· ···················· ···················· ····················
J 2497 hCoV-19/England/ALDP-6C13A2/2020|EPI_ISL_557861|20 ···················· ···················· ····················· ···················· ···················· ····················
J 2498 hCoV-19/England/ALDP-6BCB75/2020|EPI_ISL_557862|20 ···················· ···················· ····················· ···················· ···················· ····················
J 2499 hCoV-19/England/ALDP-6B556F/2020|EPI_ISL_557863|20 ···················· ···················· ····················· ···················· ···················· ····················
J 2500 hCoV-19/England/MILK-60D8C9/2020|EPI_ISL_557866|20 ···················· ···················· ····················· ···················· ···················· ····················
J 2501 hCoV-19/England/MILK-6238FA/2020|EPI_ISL_557867|20 ···················· ···················· ····················· ···················· ···················· ····················
J 2502 hCoV-19/England/MILK-63A799/2020|EPI_ISL_557869|20 ···················· ···················· ····················· ···················· ···················· ····················
J 2503 hCoV-19/England/MILK-62BC41/2020|EPI_ISL_557870|20 ···················· ···················· ····················· ···················· ···················· ····················
J 2504 hCoV-19/England/ALDP-6C3030/2020|EPI_ISL_557872|20 ···················· ···················· ····················· ···················· ···················· ····················
J 2505 hCoV-19/England/ALDP-6BEB73/2020|EPI_ISL_557873|20 ···················· ···················· ····················· ···················· ···················· ····················
J 2506 hCoV-19/England/ALDP-6B8EAD/2020|EPI_ISL_557874|20 ···················· ···················· ····················· ···················· ···················· ····················
J 2507 hCoV-19/England/ALDP-6B5ABB/2020|EPI_ISL_557875|20 ···················· ···················· ····················· ···················· ···················· ····················
J 2508 hCoV-19/England/MILK-61B2F7/2020|EPI_ISL_557876|20 ···················· ···················· ····················· ···················· ···················· ····················
J 2509 hCoV-19/England/MILK-62C457/2020|EPI_ISL_557877|20 ···················· ···················· ····················· ···················· ···················· ····················
J 2510 hCoV-19/England/MILK-639C22/2020|EPI_ISL_557878|20 ···················· ···················· ····················· ···················· ···················· ····················
J 2511 hCoV-19/England/MILK-628F78/2020|EPI_ISL_557879|20 ···················· ···················· ····················· ···················· ···················· ····················
J 2512 hCoV-19/England/ALDP-6B714B/2020|EPI_ISL_557887|20 ···················· ···················· ····················· ···················· ···················· ····················
J 2513 hCoV-19/England/ALDP-6AE174/2020|EPI_ISL_557888|20 ···················· ·A·················· ····················· ···················· ···················· ····················
J 2514 hCoV-19/England/MILK-63A025/2020|EPI_ISL_557890|20 ···················· ···················· ····················· ···················· ···················· ····················
J 2515 hCoV-19/England/ALDP-6C19FB/2020|EPI_ISL_557891|20 ···················· ···················· ····················· ···················· ···················· ····················
J 2516 hCoV-19/England/ALDP-6B58B1/2020|EPI_ISL_557892|20 ···················· ···················· ····················· ···················· ···················· ····················
J 2517 hCoV-19/England/MILK-62B50A/2020|EPI_ISL_557893|20 ···················· ···················· ····················· ···················· ···················· ····················
J 2518 hCoV-19/England/MILK-622381/2020|EPI_ISL_557894|20 ···················· ···················· ····················· ···················· ···················· ····················
J 2519 hCoV-19/England/MILK-622B4D/2020|EPI_ISL_557895|20 ···················· ···················· ····················· ···················· ···················· ····················
J 2520 hCoV-19/England/ALDP-6BE28D/2020|EPI_ISL_557897|20 ···················· ···················· ····················· ···················· ···················· ····················
J 2521 hCoV-19/England/ALDP-6C02E2/2020|EPI_ISL_557901|20 ···················· ···················· ····················· ···················· ···················· ····················
J 2522 hCoV-19/England/ALDP-6B0898/2020|EPI_ISL_557903|20 ···················· ···················· ····················· ···················· ···················· ····················
J 2523 hCoV-19/England/ALDP-6B5848/2020|EPI_ISL_557904|20 ···················· ···················· ····················· ···················· ···················· ····················
J 2524 hCoV-19/England/MILK-60A7DE/2020|EPI_ISL_557905|20 ···················· ···················· ····················· ···················· ···················· ····················
J 2525 hCoV-19/England/ALDP-6B8C3A/2020|EPI_ISL_557906|20 ···················· ···················· ····················· ···················· ···················· ····················
J 2526 hCoV-19/England/MILK-6293A8/2020|EPI_ISL_557909|20 ···················· ···················· ····················· ···················· ···················· ····················
J 2527 hCoV-19/England/MILK-61DA0C/2020|EPI_ISL_557911|20 ···················· ···················· ····················· ···················· ···················· ····················
J 2528 hCoV-19/England/ALDP-6BC3C8/2020|EPI_ISL_557912|20 ···················· ···················· ····················· ···················· ···················· ····················
J 2529 hCoV-19/England/ALDP-6B7600/2020|EPI_ISL_557914|20 ···················· ···················· ····················· ···················· ···················· ····················
J 2530 hCoV-19/England/ALDP-6C1014/2020|EPI_ISL_557916|20 ···················· ···················· ····················· ···················· ···················· ····················
J 2531 hCoV-19/England/MILK-63BE2A/2020|EPI_ISL_557919|20 ···················· ···················· ····················· ···················· ···················· ····················
J 2532 hCoV-19/England/ALDP-6B7002/2020|EPI_ISL_557920|20 ···················· ···················· ····················· ···················· ···················· ····················
J 2533 hCoV-19/England/ALDP-6AECDD/2020|EPI_ISL_557922|20 ···················· ···················· ····················· ···················· ···················· ····················
J 2534 hCoV-19/England/ALDP-6C1409/2020|EPI_ISL_557923|20 ···················· ···················· ····················· ···················· ···················· ····················
J 2535 hCoV-19/England/ALDP-6AD904/2020|EPI_ISL_557924|20 ···················· ···················· ····················· ···················· ···················· ····················
J 2536 hCoV-19/England/MILK-628312/2020|EPI_ISL_557925|20 ···················· ···················· ····················· ···················· ···················· ····················

J 2537 hCoV-19/England/MILK-60EB38/2020|EPI_ISL_557926|20 ···················· ···················· ···················· ···················· ···················· ····················
J 2538 hCoV-19/England/MILK-6160F2/2020|EPI_ISL_557927|20 ···················· ·A·················· ···················· ···················· ···················· ···A················
J 2539 hCoV-19/England/ALDP-6B4E47/2020|EPI_ISL_557928|20 ···················· ···················· ···················· ···················· ···················· ····················
J 2540 hCoV-19/England/ALDP-6BB226/2020|EPI_ISL_557929|20 ···················· ···················· ···················· ···················· ···················· ····················
J 2541 hCoV-19/England/ALDP-6BD52B/2020|EPI_ISL_557930|20 ···················· ···················· ···················· ···················· ···················· ····················
J 2542 hCoV-19/England/ALDP-6BBD9E/2020|EPI_ISL_557931|20 ···················· ···················· ···················· ···················· ···················· ····················
J 2543 hCoV-19/England/ALDP-6B5D76/2020|EPI_ISL_557932|20 ···················· ···················· ···················· ···················· ···················· ····················
J 2544 hCoV-19/England/ALDP-6B663E/2020|EPI_ISL_557934|20 ···················· ·A·················· ···················· ···················· ···················· ····················
J 2545 hCoV-19/England/ALDP-6B9FF4/2020|EPI_ISL_557935|20 ···················· ···················· ···················· ···················· ···················· ····················
J 2546 hCoV-19/England/MILK-62104F/2020|EPI_ISL_557938|20 ···················· ···················· ···················· ···················· ···················· ····················
J 2547 hCoV-19/England/ALDP-6BB5D2/2020|EPI_ISL_557939|20 ···················· ···················· ···················· ···················· ···················· ····················
J 2548 hCoV-19/England/MILK-636663/2020|EPI_ISL_557940|20 ···················· ···················· ···················· ···················· ···················· ····················
J 2549 hCoV-19/England/MILK-627CFA/2020|EPI_ISL_557941|20 ···········T········ ···················· ···················· ···················· ···················· ····················
J 2550 hCoV-19/England/MILK-60C8BB/2020|EPI_ISL_557942|20 ···················· ···················· ···················· ···················· ···················· ····················
J 2551 hCoV-19/England/ALDP-6B7B6B/2020|EPI_ISL_557943|20 ···················· ···················· ···················· ···················· ···················· ····················
J 2552 hCoV-19/England/MILK-62C40C/2020|EPI_ISL_557944|20 ···················· ···················· ···················· ···················· ···················· ····················
J 2553 hCoV-19/Wales/ALDP-6B2EA3/2020|EPI_ISL_557945|2020 ···················· ···················· ···················· ···················· ···················· ····················
J 2554 hCoV-19/Wales/ALDP-6B2FA0/2020|EPI_ISL_557946|2020 ···················· ···················· ···················· ···················· ···················· ····················
J 2555 hCoV-19/England/MILK-62ACF7/2020|EPI_ISL_557947|20 ···················· ···················· ···················· ···················· ···················· ····················
J 2556 hCoV-19/England/MILK-623809/2020|EPI_ISL_557949|20 ···················· ···················· ···················· ···················· ···················· ····················
J 2557 hCoV-19/England/ALDP-6B9FB8/2020|EPI_ISL_557950|20 ···················· ···················· ···················· ···················· ···················· ····················
J 2558 hCoV-19/England/ALDP-6BAC0B/2020|EPI_ISL_557951|20 ···················· ···················· ···················· ···················· ···················· ····················
J 2559 hCoV-19/England/MILK-62D81E/2020|EPI_ISL_557953|20 ···················· ···················· ···················· ···················· ···················· ····················
J 2560 hCoV-19/England/MILK-62CD98/2020|EPI_ISL_557954|20 ···················· ···················· ···················· ···················· ···················· ····················
J 2561 hCoV-19/England/MILK-9A899D/2020|EPI_ISL_549437|20 ···················· ···················· ···················· ···················· ···················· ····················
J 2562 hCoV-19/England/MILK-9A8A03/2020|EPI_ISL_549438|20 ···················· ···················· ···················· ···················· ···················· ····················
J 2563 hCoV-19/England/MILK-9A8A5E/2020|EPI_ISL_549440|20 ···················· ···················· ···················· ···················· ···················· ····················
J 2564 hCoV-19/England/MILK-9A8A6D/2020|EPI_ISL_549441|20 ···················· ·A·················· ···················· ···················· ···················· ····················
J 2565 hCoV-19/England/MILK-9AB569/2020|EPI_ISL_549442|20 ···················· ···················· ···················· ···················· ···················· ····················
J 2566 hCoV-19/England/MILK-9AB578/2020|EPI_ISL_549443|20 ···················· ···················· ···················· ···················· ···················· ····················
J 2567 hCoV-19/England/MILK-9AB596/2020|EPI_ISL_549444|20 ···················· ···················· ···················· ···················· ···················· ····················
J 2568 hCoV-19/England/MILK-9AB5A5/2020|EPI_ISL_549445|20 ···················· ···················· ···················· ···················· ···················· ····················
J 2569 hCoV-19/England/MILK-9AB5B4/2020|EPI_ISL_549446|20 ···················· ···················· ···················· ···················· ···················· ····················
J 2570 hCoV-19/England/MILK-9AB5C3/2020|EPI_ISL_549447|20 ···················· ···················· ···················· ···················· ···················· ····················
J 2571 hCoV-19/England/MILK-9AB60C/2020|EPI_ISL_549448|20 ···················· ···················· ···················· ···················· ···················· ····················
J 2572 hCoV-19/England/MILK-9AB61B/2020|EPI_ISL_549449|20 ···················· ···················· ···················· ···················· ···················· ····················
J 2573 hCoV-19/England/MILK-9AB648/2020|EPI_ISL_549450|20 ···················· ···················· ···················· ···················· ···················· ····················
J 2574 hCoV-19/England/MILK-9AB657/2020|EPI_ISL_549451|20 ···················· ···················· ···················· ···················· ···················· ····················
J 2575 hCoV-19/England/MILK-9AB666/2020|EPI_ISL_549452|20 ···················· ···················· ···················· ···················· ···················· ····················
J 2576 hCoV-19/England/MILK-9AB675/2020|EPI_ISL_549453|20 ···················· ···················· ···················· ···················· ···················· ····················
J 2577 hCoV-19/England/MILK-9AB6DF/2020|EPI_ISL_549454|20 ···················· ···················· ···················· ···················· ···················· ····················
J 2578 hCoV-19/England/MILK-9AB709/2020|EPI_ISL_549455|20 ···················· ···················· ···················· ···················· ···················· ····················
J 2579 hCoV-19/England/MILK-9AB718/2020|EPI_ISL_549456|20 ···················· ···················· ···················· ···················· ···················· ····················
J 2580 hCoV-19/England/MILK-9AB745/2020|EPI_ISL_549457|20 ···················· ···················· ···················· ···················· ···················· ····················
J 2581 hCoV-19/England/MILK-9AB772/2020|EPI_ISL_549458|20 ···················· ···················· ···················· ···················· ···················· ····················
J 2582 hCoV-19/England/MILK-9AB781/2020|EPI_ISL_549459|20 ···················· ···················· ···················· ···················· ···················· ····················
J 2583 hCoV-19/England/MILK-9C4676/2020|EPI_ISL_549460|20 ···················· ···················· ···················· ···················· ···················· ····················
J 2584 hCoV-19/England/MILK-9C4773/2020|EPI_ISL_549461|20 ···················· ···················· ···················· ···················· ···················· ····················
J 2585 hCoV-19/England/MILK-9C4782/2020|EPI_ISL_549462|20 ···················· ···················· ···················· ···················· ···················· ····················
J 2586 hCoV-19/England/MILK-9C47A0/2020|EPI_ISL_549463|20 ···················· ···················· ···················· ···················· ···················· ····················
J 2587 hCoV-19/England/MILK-9C47BF/2020|EPI_ISL_549464|20 ···················· ·A·················· ···················· ···················· ···················· ···A················
J 2588 hCoV-19/England/MILK-9C47EC/2020|EPI_ISL_549465|20 ···················· ·A·················· ···················· ···················· ···················· ···A················
J 2589 hCoV-19/England/MILK-9C4807/2020|EPI_ISL_549466|20 ···················· ···················· ···················· ···················· ···················· ····················
J 2590 hCoV-19/England/MILK-9C4825/2020|EPI_ISL_549467|20 ···················· ···················· ···················· ···················· ···················· ····················
J 2591 hCoV-19/England/MILK-9C4834/2020|EPI_ISL_549468|20 ···················· ···················· ···················· ···················· ···················· ····················
J 2592 hCoV-19/England/MILK-9C4843/2020|EPI_ISL_549469|20 ···················· ·A·················· ···················· ···················· ···················· ····················
J 2593 hCoV-19/England/MILK-9C4852/2020|EPI_ISL_549470|20 ···················· ···················· ···················· ···················· ···················· ····················
J 2594 hCoV-19/England/MILK-9C498C/2020|EPI_ISL_549471|20 ···················· ···················· ···················· ···················· ···················· ····················
J 2595 hCoV-19/England/MILK-9C49AA/2020|EPI_ISL_549472|20 ···················· ····A··············· ···················· ···················· ···················· ····················
J 2596 hCoV-19/England/MILK-9C4A01/2020|EPI_ISL_549473|20 ···················· ·A·················· ···················· ···················· ···················· ···A················
J 2597 hCoV-19/England/MILK-9C4A2F/2020|EPI_ISL_549474|20 ···················· ···················· ···················· ···················· ···················· ····················
J 2598 hCoV-19/England/MILK-9C4A6B/2020|EPI_ISL_549475|20 ···················· ···················· ···················· ···················· ···················· ····················
J 2599 hCoV-19/England/MILK-9C4AB6/2020|EPI_ISL_549476|20 ···················· ···················· ···················· ···················· ···················· ····················
J 2600 hCoV-19/England/NORW-EE651/2020|EPI_ISL_549477|202 ···················· ···················· ···················· ···················· ···················· ····················
J 2601 hCoV-19/England/NORW-EE6D9/2020|EPI_ISL_549478|202 ···················· ···················· ···················· ···················· ···················· ····················
J 2602 hCoV-19/England/NORW-EF5BD/2020|EPI_ISL_549479|202 ···················· ···················· ···················· ···················· ···················· ····················
J 2603 hCoV-19/England/NORW-EF869/2020|EPI_ISL_549480|202 ···················· ···················· ···················· ···················· ···················· ····················
J 2604 hCoV-19/England/NORW-EFA09/2020|EPI_ISL_549481|202 ···················· ···················· ···················· ···················· ···················· ····················
J 2605 hCoV-19/England/NORW-EFA27/2020|EPI_ISL_549482|202 ···················· ···················· ···················· ···················· ···················· ····················
J 2606 hCoV-19/England/NORW-EFACD/2020|EPI_ISL_549483|202 ···················· ···················· ···················· ···················· ···················· ····················
J 2607 hCoV-19/England/NORW-EFADC/2020|EPI_ISL_549484|202 ···················· ···················· ···················· ···················· ···················· ····················
J 2608 hCoV-19/England/NORW-EFB06/2020|EPI_ISL_549485|202 ···················· ···················· ···················· ···················· ···················· ····················
J 2609 hCoV-19/England/NORW-EFB60/2020|EPI_ISL_549486|202 ···················· ···················· ···················· ···················· ···················· ····················
J 2610 hCoV-19/England/NORW-EFC7C/2020|EPI_ISL_549487|202 ···················· ···················· ···················· ···················· ···················· ····················
J 2611 hCoV-19/England/NORW-EFC8B/2020|EPI_ISL_549488|202 ···················· ···················· ···················· ···················· ···················· ····················
J 2612 hCoV-19/England/NORW-EFCD6/2020|EPI_ISL_549489|202 ···················· ···················· ···················· ···················· ···················· ····················
J 2613 hCoV-19/England/NORW-EFCE5/2020|EPI_ISL_549490|202 ···················· ···················· ···················· ···················· ···················· ····················
J 2614 hCoV-19/England/NOTT-11329A/2020|EPI_ISL_549491|20 ···················· ···················· ···················· ···················· ···················· ····················
J 2615 hCoV-19/England/NOTT-1132B8/2020|EPI_ISL_549492|20 ···················· ·A·················· ···················· ···················· ···················· ····················
J 2616 hCoV-19/England/NOTT-1132C7/2020|EPI_ISL_549493|20 ···················· ···················· ···················· ···················· ···················· ····················

| | | | | | | | |
|---|---|---|---|---|---|---|---|
| J | 2617 hCoV-19/England/NOTT-1132E5/2020\|EPI_ISL_549494\|20 | ···················· | ···················· | ····················· | ···················· | ···················· | ···················· |
| J | 2618 hCoV-19/England/NOTT-1132F4/2020\|EPI_ISL_549495\|20 | ···················· | ···················· | ····················· | ···················· | ···················· | ···················· |
| J | 2619 hCoV-19/England/NOTT-113300/2020\|EPI_ISL_549496\|20 | ···················· | ···················· | ····················· | ···················· | ···················· | ···················· |
| J | 2620 hCoV-19/England/NOTT-11333D/2020\|EPI_ISL_549497\|20 | ···················· | ·A·················· | ····················· | ···················· | ···················· | ···················· |
| J | 2621 hCoV-19/England/NOTT-11334C/2020\|EPI_ISL_549498\|20 | ···················· | ·A·················· | ····················· | ···················· | ···················· | ···················· |
| J | 2622 hCoV-19/England/NOTT-11335B/2020\|EPI_ISL_549499\|20 | ···················· | ···················· | ····················· | ···················· | ···················· | ···················· |
| J | 2623 hCoV-19/England/MILK-99A2EE/2020\|EPI_ISL_550410\|20 | ···················· | ···················· | ····················· | ···················· | ···················· | ···················· |
| J | 2624 hCoV-19/England/MILK-999D75/2020\|EPI_ISL_550412\|20 | ···················· | ···················· | ····················· | ···················· | ···················· | ···················· |
| J | 2625 hCoV-19/England/MILK-99BAE5/2020\|EPI_ISL_550414\|20 | ···················· | ···················· | ····················· | ···················· | ···················· | ···················· |
| J | 2626 hCoV-19/England/MILK-99A6B5/2020\|EPI_ISL_550415\|20 | ···················· | ···················· | ····················· | ···················· | ···················· | ···················· |
| J | 2627 hCoV-19/England/MILK-977D22/2020\|EPI_ISL_550417\|20 | ···················· | ·A·················· | ····················· | ···················· | ···················· | ··A················· |
| J | 2628 hCoV-19/England/MILK-9773E4/2020\|EPI_ISL_550418\|20 | ···················· | ···················· | ····················· | ···················· | ···················· | ···················· |
| J | 2629 hCoV-19/England/MILK-9787C9/2020\|EPI_ISL_550419\|20 | ···················· | ···················· | ····················· | ···················· | ···················· | ···················· |
| J | 2630 hCoV-19/England/MILK-977575/2020\|EPI_ISL_550420\|20 | ···················· | ···················· | ···········T········· | ···················· | ···················· | ···················· |
| J | 2631 hCoV-19/Wales/MILK-9789C3/2020\|EPI_ISL_550421\|2020 | ···················· | ···················· | ····················· | ···················· | ···················· | ···················· |
| J | 2632 hCoV-19/England/MILK-977593/2020\|EPI_ISL_550422\|20 | ···················· | ·A·················· | ····················· | ···················· | ···················· | ··A················· |
| J | 2633 hCoV-19/England/MILK-97892D/2020\|EPI_ISL_550423\|20 | ···················· | ·A·················· | ····················· | ···················· | ···················· | ··A················· |
| J | 2634 hCoV-19/England/MILK-978750/2020\|EPI_ISL_550428\|20 | ···················· | ·A·················· | ····················· | ···················· | ···················· | ··A················· |
| J | 2635 hCoV-19/England/MILK-9776EB/2020\|EPI_ISL_550430\|20 | ···················· | ···················· | ····················· | ···················· | ···················· | ···················· |
| J | 2636 hCoV-19/England/MILK-97741E/2020\|EPI_ISL_550431\|20 | ···················· | ···················· | ····················· | ···················· | ···················· | ···················· |
| J | 2637 hCoV-19/England/MILK-9780CE/2020\|EPI_ISL_550432\|20 | ···················· | ···················· | ····················· | ···················· | ···················· | ···················· |
| J | 2638 hCoV-19/England/MILK-978608/2020\|EPI_ISL_550434\|20 | ···················· | ···················· | ····················· | ···················· | ···················· | ···················· |
| J | 2639 hCoV-19/England/MILK-977566/2020\|EPI_ISL_550435\|20 | ···················· | ···················· | ····················· | ···················· | ···················· | ···················· |
| J | 2640 hCoV-19/England/MILK-9773B7/2020\|EPI_ISL_550437\|20 | ···················· | ···················· | ····················· | ···················· | ···················· | ···················· |
| J | 2641 hCoV-19/England/MILK-9789A5/2020\|EPI_ISL_550438\|20 | ···················· | ···················· | ····················· | ···················· | ···················· | ···················· |
| J | 2642 hCoV-19/England/MILK-97893C/2020\|EPI_ISL_550439\|20 | ···················· | ···················· | ····················· | ···················· | ···················· | ···················· |
| J | 2643 hCoV-19/England/MILK-9785ED/2020\|EPI_ISL_550440\|20 | ···················· | ···················· | ····················· | ···················· | ···················· | ···················· |
| J | 2644 hCoV-19/England/MILK-9777E8/2020\|EPI_ISL_550442\|20 | ···················· | ···················· | ····················· | ···················· | ···················· | ···················· |
| J | 2645 hCoV-19/England/MILK-977C34/2020\|EPI_ISL_550443\|20 | ···················· | ···················· | ····················· | ···················· | ···················· | ···················· |
| J | 2646 hCoV-19/England/MILK-9781AD/2020\|EPI_ISL_550445\|20 | ···················· | ···················· | ····················· | ···················· | ···················· | ···················· |
| J | 2647 hCoV-19/England/MILK-977FA4/2020\|EPI_ISL_550446\|20 | ···················· | ···················· | ····················· | ···················· | ···················· | ···················· |
| J | 2648 hCoV-19/England/MILK-977469/2020\|EPI_ISL_550447\|20 | ···················· | ···················· | ····················· | ···················· | ···················· | ···················· |
| J | 2649 hCoV-19/England/MILK-978AFD/2020\|EPI_ISL_550448\|20 | ···················· | ·A·················· | ····················· | ···················· | ···················· | ··A················· |
| J | 2650 hCoV-19/England/MILK-978671/2020\|EPI_ISL_550449\|20 | ···················· | ···················· | ····················· | ···················· | ···················· | ···················· |
| J | 2651 hCoV-19/England/MILK-9780DD/2020\|EPI_ISL_550450\|20 | ···················· | ···················· | ····················· | ···················· | ···················· | ···················· |
| J | 2652 hCoV-19/England/MILK-977F59/2020\|EPI_ISL_550451\|20 | ···················· | ···················· | ····················· | ···················· | ···················· | ···················· |
| J | 2653 hCoV-19/England/MILK-97733F/2020\|EPI_ISL_550452\|20 | ···················· | ···················· | ····················· | ···················· | ···················· | ···················· |
| J | 2654 hCoV-19/England/MILK-978AEE/2020\|EPI_ISL_550453\|20 | ···················· | ·A·················· | ····················· | ···················· | ···················· | ··A················· |
| J | 2655 hCoV-19/England/MILK-97779D/2020\|EPI_ISL_550454\|20 | ···················· | ·A·················· | ····················· | ···················· | ···················· | ···················· |
| J | 2656 hCoV-19/England/MILK-978152/2020\|EPI_ISL_550456\|20 | ···················· | ···················· | ····················· | ···················· | ···················· | ···················· |
| J | 2657 hCoV-19/England/MILK-978741/2020\|EPI_ISL_550457\|20 | ···················· | ···················· | ····················· | ···················· | ···················· | ···················· |
| J | 2658 hCoV-19/Wales/MILK-9775C0/2020\|EPI_ISL_550458\|2020 | ···················· | ···················· | ····················· | ···················· | ···················· | ···················· |
| J | 2659 hCoV-19/England/MILK-97737B/2020\|EPI_ISL_550459\|20 | ···················· | ···················· | ····················· | ···················· | ···················· | ···················· |
| J | 2660 hCoV-19/England/MILK-97876F/2020\|EPI_ISL_550460\|20 | ···················· | ·A·················· | ····················· | ···················· | ···················· | ··A················· |
| J | 2661 hCoV-19/England/MILK-978107/2020\|EPI_ISL_550461\|20 | ···················· | ···················· | ····················· | ···················· | ···················· | ···················· |
| J | 2662 hCoV-19/England/MILK-977627/2020\|EPI_ISL_550462\|20 | ···················· | ···················· | ····················· | ···················· | ···················· | ···················· |
| J | 2663 hCoV-19/England/MILK-97869F/2020\|EPI_ISL_550463\|20 | ···················· | ···················· | ····················· | ···················· | ···················· | ···················· |
| J | 2664 hCoV-19/England/MILK-97752A/2020\|EPI_ISL_550464\|20 | ···················· | ···················· | ····················· | ···················· | ···················· | ···················· |
| J | 2665 hCoV-19/England/MILK-9780FB/2020\|EPI_ISL_550465\|20 | ···················· | ···················· | ····················· | ···················· | ···················· | ···················· |
| J | 2666 hCoV-19/England/MILK-977979/2020\|EPI_ISL_550466\|20 | ···················· | ···················· | ····················· | ···················· | ···················· | ···················· |
| J | 2667 hCoV-19/England/MILK-978A93/2020\|EPI_ISL_550468\|20 | ···················· | ···················· | ····················· | ···················· | ···················· | ···················· |
| J | 2668 hCoV-19/England/MILK-977F77/2020\|EPI_ISL_550469\|20 | ···················· | ···················· | ····················· | ···················· | ···················· | ···················· |
| J | 2669 hCoV-19/England/MILK-97894B/2020\|EPI_ISL_550470\|20 | ···················· | ···················· | ····················· | ···················· | ···················· | ···················· |
| J | 2670 hCoV-19/England/MILK-9779A6/2020\|EPI_ISL_550471\|20 | ···················· | ···················· | ····················· | ···················· | ···················· | ···················· |
| J | 2671 hCoV-19/England/MILK-977D04/2020\|EPI_ISL_550472\|20 | ···················· | ·············A······ | ····················· | ···················· | ···················· | ···················· |
| J | 2672 hCoV-19/England/MILK-9777BB/2020\|EPI_ISL_550473\|20 | ···················· | ···················· | ····················· | ···················· | ···················· | ···················· |
| J | 2673 hCoV-19/England/MILK-978653/2020\|EPI_ISL_550475\|20 | ···················· | ···················· | ···········T········· | ···················· | ···················· | ···················· |
| J | 2674 hCoV-19/England/MILK-978A0C/2020\|EPI_ISL_550478\|20 | ···················· | ···················· | ····················· | ···················· | ···················· | ···················· |
| J | 2675 hCoV-19/England/MILK-97794C/2020\|EPI_ISL_550479\|20 | ···················· | ···················· | ····················· | ···················· | ···················· | ···················· |
| J | 2676 hCoV-19/England/MILK-977548/2020\|EPI_ISL_550480\|20 | ···················· | ···················· | ····················· | ···················· | ···················· | ···················· |
| J | 2677 hCoV-19/England/MILK-978046/2020\|EPI_ISL_550481\|20 | ···················· | ···················· | ····················· | ···················· | ···················· | ···················· |
| J | 2678 hCoV-19/England/MILK-977E01/2020\|EPI_ISL_550483\|20 | ···················· | ···················· | ····················· | ···················· | ···················· | ···················· |
| J | 2679 hCoV-19/England/ALDP-684B94/2020\|EPI_ISL_551740\|20 | ···················· | ···················· | ····················· | ···················· | ···················· | ···················· |
| J | 2680 hCoV-19/England/MILK-6A759D/2020\|EPI_ISL_551741\|20 | ···················· | ···················· | ····················· | ···················· | ···················· | ···················· |
| J | 2681 hCoV-19/England/ALDP-688DA9/2020\|EPI_ISL_551742\|20 | ···················· | ···················· | ····················· | ···················· | ···················· | ···················· |
| J | 2682 hCoV-19/England/ALDP-687EB6/2020\|EPI_ISL_551743\|20 | ···················· | ···················· | ····················· | ···················· | ·····C·············· | ···················· |
| J | 2683 hCoV-19/England/MILK-699FBE/2020\|EPI_ISL_551744\|20 | ···················· | ···················· | ····················· | ···················· | ···················· | ···················· |
| J | 2684 hCoV-19/England/MILK-6A5BBB/2020\|EPI_ISL_551745\|20 | ···················· | ···················· | ····················· | ···················· | ···················· | ···················· |
| J | 2685 hCoV-19/England/MILK-6A027F/2020\|EPI_ISL_551747\|20 | ···················· | ···················· | ····················· | ···················· | ···················· | ···················· |
| J | 2686 hCoV-19/England/MILK-69C0B8/2020\|EPI_ISL_551748\|20 | ···················· | ···················· | ····················· | ···················· | ···················· | ···················· |
| J | 2687 hCoV-19/England/ALDP-682CB1/2020\|EPI_ISL_551749\|20 | ···················· | ···················· | ····················· | ···················· | ···················· | ···················· |
| J | 2688 hCoV-19/England/MILK-6A51D7/2020\|EPI_ISL_551752\|20 | ···················· | ···················· | ····················· | ···················· | ···················· | ···················· |
| J | 2689 hCoV-19/England/MILK-6AB0B6/2020\|EPI_ISL_551753\|20 | ···················· | ···················· | ····················· | ···················· | ···················· | ···················· |
| J | 2690 hCoV-19/England/MILK-6798D7/2020\|EPI_ISL_551754\|20 | ···················· | ···················· | ···········T········· | ···················· | ···················· | ···················· |
| J | 2691 hCoV-19/England/ALDP-68397D/2020\|EPI_ISL_551755\|20 | ···················· | ···················· | ····················· | ···················· | ···················· | ···················· |
| J | 2692 hCoV-19/England/MILK-69CB3F/2020\|EPI_ISL_551758\|20 | ···················· | ···················· | ····················· | ···················· | ···················· | ···················· |
| J | 2693 hCoV-19/England/ALDP-6898E3/2020\|EPI_ISL_551759\|20 | ···················· | ···················· | ····················· | ···················· | ···················· | ···················· |
| J | 2694 hCoV-19/Wales/ALDP-689E5A/2020\|EPI_ISL_551760\|2020 | ···················· | ···················· | ····················· | ···················· | ···················· | ···················· |
| J | 2695 hCoV-19/England/ALDP-68844A/2020\|EPI_ISL_551761\|20 | ···················· | ···················· | ····················· | ···················· | ···················· | ···················· |
| J | 2696 hCoV-19/England/ALDP-689810/2020\|EPI_ISL_551762\|20 | ···················· | ···················· | ····················· | ···················· | ···················· | ···················· |

| | | | | | | | |
|---|---|---|---|---|---|---|---|
| J | 2697 hCoV-19/England/ALDP-683852/2020\|EPI_ISL_551763\|20 | .................... | .................... | .................... | .................... | .................... | .................... |
| J | 2698 hCoV-19/England/ALDP-67901B/2020\|EPI_ISL_551764\|20 | .................... | .................... | .................... | .................... | .................... | .................... |
| J | 2699 hCoV-19/England/MILK-69F194/2020\|EPI_ISL_551765\|20 | .................... | .................... | .................... | .................... | .................... | .................... |
| J | 2700 hCoV-19/England/MILK-6A2969/2020\|EPI_ISL_551766\|20 | .................... | .................... | .................... | .................... | .................... | .................... |
| J | 2701 hCoV-19/England/ALDP-67E09E/2020\|EPI_ISL_551772\|20 | .................... | .................... | .................... | .................... | .................... | .................... |
| J | 2702 hCoV-19/England/MILK-67C5FB/2020\|EPI_ISL_551773\|20 | .................... | .................... | .................... | .................... | .................... | .................... |
| J | 2703 hCoV-19/England/MILK-6A8E64/2020\|EPI_ISL_551774\|20 | .................... | .................... | .................... | .................... | .................... | .................... |
| J | 2704 hCoV-19/England/ALDP-68696B/2020\|EPI_ISL_551776\|20 | .................... | .................... | .................... | .................... | .................... | .................... |
| J | 2705 hCoV-19/Wales/MILK-6AA406/2020\|EPI_ISL_551777\|2020 | .................... | .................... | .................... | .................... | .................... | .................... |
| J | 2706 hCoV-19/England/ALDP-684208/2020\|EPI_ISL_551779\|20 | .................... | .................... | .................... | .................... | .................... | .................... |
| J | 2707 hCoV-19/England/ALDP-6789B7/2020\|EPI_ISL_551780\|20 | .................... | .................... | .................... | .................... | .................... | .................... |
| J | 2708 hCoV-19/England/MILK-696BEA/2020\|EPI_ISL_551781\|20 | .................... | .................... | .................... | .................... | .................... | .................... |
| J | 2709 hCoV-19/England/MILK-6A294B/2020\|EPI_ISL_551782\|20 | .................... | .................... | .................... | .................... | .................... | .................... |
| J | 2710 hCoV-19/England/ALDP-689AFC/2020\|EPI_ISL_551784\|20 | .................... | .................... | .................... | .................... | .................... | .................... |
| J | 2711 hCoV-19/England/ALDP-683D08/2020\|EPI_ISL_551785\|20 | .................... | .................... | .................... | .................... | .................... | .................... |
| J | 2712 hCoV-19/England/MILK-6938B9/2020\|EPI_ISL_551786\|20 | .................... | .................... | .................... | .................... | .................... | .................... |
| J | 2713 hCoV-19/England/ALDP-67D2B7/2020\|EPI_ISL_551789\|20 | .................... | .................... | .................... | .................... | .................... | .................... |
| J | 2714 hCoV-19/England/MILK-6A69DE/2020\|EPI_ISL_551790\|20 | .................... | .................... | .................... | .................... | .................... | .................... |
| J | 2715 hCoV-19/England/MILK-6A185E/2020\|EPI_ISL_551791\|20 | .................... | .................... | .................... | .................... | .................... | .................... |
| J | 2716 hCoV-19/England/MILK-69DA31/2020\|EPI_ISL_551792\|20 | .................... | .................... | .................... | .................... | .................... | .................... |
| J | 2717 hCoV-19/England/MILK-6A7E47/2020\|EPI_ISL_551793\|20 | .................... | .................... | .................... | .................... | .................... | .................... |
| J | 2718 hCoV-19/England/MILK-6A0C80/2020\|EPI_ISL_551794\|20 | .................... | .................... | .................... | .................... | .................... | .................... |
| J | 2719 hCoV-19/England/ALDP-6839B9/2020\|EPI_ISL_551795\|20 | .................... | .................... | .................... | .................... | .................... | .................... |
| J | 2720 hCoV-19/England/ALDP-689FB1/2020\|EPI_ISL_551796\|20 | .................... | .................... | .................... | .................... | .................... | .................... |
| J | 2721 hCoV-19/England/MILK-677903/2020\|EPI_ISL_551797\|20 | .................... | .................... | ..........T......... | .................... | .................... | .................... |
| J | 2722 hCoV-19/England/ALDP-688D03/2020\|EPI_ISL_551798\|20 | .................... | .................... | .................... | .................... | .................... | .................... |
| J | 2723 hCoV-19/England/ALDP-67D7C7/2020\|EPI_ISL_551799\|20 | .................... | .................... | .................... | .................... | .................... | .................... |
| J | 2724 hCoV-19/England/ALDP-9303D8/2020\|EPI_ISL_552901\|20 | .................... | .................... | .................... | .................... | .................... | .................... |
| J | 2725 hCoV-19/England/ALDP-935AA1/2020\|EPI_ISL_552902\|20 | .................... | .................... | .................... | .................... | .................... | .................... |
| J | 2726 hCoV-19/England/ALDP-935564/2020\|EPI_ISL_552903\|20 | .................... | .................... | .................... | .................... | .................... | .................... |
| J | 2727 hCoV-19/England/ALDP-9317F9/2020\|EPI_ISL_552904\|20 | .................... | .................... | .................... | .................... | .................... | .................... |
| J | 2728 hCoV-19/England/MILK-938EDF/2020\|EPI_ISL_552905\|20 | .................... | .................... | .................... | .................... | .................... | .................... |
| J | 2729 hCoV-19/England/ALDP-903356/2020\|EPI_ISL_552906\|20 | .................... | .................... | .................... | .................... | .................... | .................... |
| J | 2730 hCoV-19/England/ALDP-931674/2020\|EPI_ISL_552907\|20 | .................... | .................... | .................... | .................... | .................... | .................... |
| J | 2731 hCoV-19/England/ALDP-91FF62/2020\|EPI_ISL_552908\|20 | .................... | .................... | .................... | .................... | .................... | .................... |
| J | 2732 hCoV-19/England/ALDP-920DCE/2020\|EPI_ISL_552909\|20 | .................... | .A.................. | .................... | .................... | .................... | .................... |
| J | 2733 hCoV-19/England/MILK-93B73B/2020\|EPI_ISL_552910\|20 | .................... | .................... | .................... | .................... | .................... | .................... |
| J | 2734 hCoV-19/England/ALDP-9150C3/2020\|EPI_ISL_552911\|20 | .................... | .................... | .................... | .................... | .................... | .................... |
| J | 2735 hCoV-19/England/ALDP-903505/2020\|EPI_ISL_552914\|20 | .................... | .................... | .................... | .................... | .................... | .................... |
| J | 2736 hCoV-19/England/ALDP-926A94/2020\|EPI_ISL_552917\|20 | .................... | .................... | .................... | .................... | .................... | .................... |
| J | 2737 hCoV-19/England/ALDP-920A5E/2020\|EPI_ISL_552918\|20 | .................... | .A.................. | .................... | .................... | .................... | .................... |
| J | 2738 hCoV-19/England/ALDP-935E2D/2020\|EPI_ISL_552919\|20 | .................... | .A.................. | .................... | .................... | .................... | .................... |
| J | 2739 hCoV-19/England/ALDP-9500DB/2020\|EPI_ISL_555139\|20 | .................... | .................... | .................... | .................... | .................... | .................... |
| J | 2740 hCoV-19/England/ALDP-9504DF/2020\|EPI_ISL_555141\|20 | .................... | .................... | .................... | .................... | .................... | .................... |
| J | 2741 hCoV-19/England/ALDP-94DD07/2020\|EPI_ISL_555142\|20 | .................... | .................... | .................... | .................... | .................... | .................... |
| J | 2742 hCoV-19/England/ALDP-95035A/2020\|EPI_ISL_555143\|20 | .................... | .A.................. | .................... | .................... | .................... | .................... |
| J | 2743 hCoV-19/England/ALDP-94DA3D/2020\|EPI_ISL_555147\|20 | .................... | .................... | .................... | .................... | .................... | .................... |
| J | 2744 hCoV-19/Wales/ALDP-9502E4/2020\|EPI_ISL_555149\|2020 | .................... | .................... | .................... | .................... | .................... | .................... |
| J | 2745 hCoV-19/England/ALDP-94FF96/2020\|EPI_ISL_555150\|20 | .................... | .................... | .................... | .................... | .................... | .................... |
| J | 2746 hCoV-19/Wales/ALDP-9503B4/2020\|EPI_ISL_555151\|2020 | .................... | .................... | .................... | .................... | .................... | .................... |
| J | 2747 hCoV-19/England/ALDP-950299/2020\|EPI_ISL_555152\|20 | .................... | .................... | .................... | .................... | .................... | .................... |
| J | 2748 hCoV-19/England/ALDP-950396/2020\|EPI_ISL_555153\|20 | .................... | .A.................. | .................... | .................... | .................... | .................... |
| J | 2749 hCoV-19/England/ALDP-94F998/2020\|EPI_ISL_555154\|20 | .................... | .................... | .................... | .................... | .................... | .................... |
| J | 2750 hCoV-19/England/ALDP-94DA79/2020\|EPI_ISL_555155\|20 | .................... | .A.................. | .................... | .................... | .................... | .................... |
| J | 2751 hCoV-19/England/ALDP-94E243/2020\|EPI_ISL_555156\|20 | .................... | .................... | .................... | .................... | .................... | .................... |
| J | 2752 hCoV-19/England/ALDP-95018D/2020\|EPI_ISL_555157\|20 | .................... | .................... | .................... | .................... | .................... | .................... |
| J | 2753 hCoV-19/England/ALDP-94FC53/2020\|EPI_ISL_555158\|20 | .................... | .................... | .................... | .................... | .................... | .................... |
| J | 2754 hCoV-19/Wales/ALDP-9506F7/2020\|EPI_ISL_555159\|2020 | .................... | .................... | .................... | .................... | .................... | .................... |
| J | 2755 hCoV-19/England/ALDP-950572/2020\|EPI_ISL_555160\|20 | .................... | .A.................. | .................... | .................... | .................... | .................... |
| J | 2756 hCoV-19/England/ALDP-94F901/2020\|EPI_ISL_555161\|20 | .................... | .................... | .................... | .................... | .................... | .................... |
| J | 2757 hCoV-19/England/ALDP-9506BB/2020\|EPI_ISL_555162\|20 | .................... | .................... | .................... | .................... | .................... | .................... |
| J | 2758 hCoV-19/England/ALDP-94FFF0/2020\|EPI_ISL_555164\|20 | .................... | .A.................. | .................... | .................... | .................... | .................... |
| J | 2759 hCoV-19/England/ALDP-94DA4C/2020\|EPI_ISL_555165\|20 | .................... | .................... | .................... | .................... | .................... | .................... |
| J | 2760 hCoV-19/England/ALDP-950044/2020\|EPI_ISL_555166\|20 | .................... | .................... | .................... | .................... | .................... | .................... |
| J | 2761 hCoV-19/England/ALDP-94FE20/2020\|EPI_ISL_555167\|20 | .................... | .................... | .................... | .................... | .................... | .................... |
| J | 2762 hCoV-19/England/ALDP-94D9B8/2020\|EPI_ISL_555168\|20 | .................... | .................... | .................... | .................... | .................... | .................... |
| J | 2763 hCoV-19/England/ALDP-950536/2020\|EPI_ISL_555169\|20 | .................... | .................... | .................... | .................... | .................... | .................... |
| J | 2764 hCoV-19/England/ALDP-94FE7B/2020\|EPI_ISL_555170\|20 | .................... | .................... | .................... | .................... | .................... | .................... |
| J | 2765 hCoV-19/England/ALDP-94FA59/2020\|EPI_ISL_555171\|20 | .................... | .................... | .................... | .................... | .................... | .................... |
| J | 2766 hCoV-19/England/ALDP-94E322/2020\|EPI_ISL_555172\|20 | .................... | .................... | .................... | .................... | .................... | .................... |
| J | 2767 hCoV-19/England/ALDP-950493/2020\|EPI_ISL_555174\|20 | .................... | .................... | .................... | .................... | .................... | .................... |
| J | 2768 hCoV-19/England/ALDP-94E155/2020\|EPI_ISL_555175\|20 | .................... | .................... | .................... | .................... | .................... | .................... |
| J | 2769 hCoV-19/England/ALDP-94FA77/2020\|EPI_ISL_555176\|20 | .................... | .................... | .................... | .................... | .................... | .................... |
| J | 2770 hCoV-19/England/ALDP-950581/2020\|EPI_ISL_555177\|20 | .................... | .................... | .................... | .................... | .................... | .................... |
| J | 2771 hCoV-19/England/ALDP-94DED7/2020\|EPI_ISL_555179\|20 | .................... | .................... | .................... | .................... | .................... | .................... |
| J | 2772 hCoV-19/England/ALDP-94FC26/2020\|EPI_ISL_555180\|20 | .................... | .................... | .................... | .................... | .................... | .................... |
| J | 2773 hCoV-19/England/ALDP-94E708/2020\|EPI_ISL_555181\|20 | .................... | .................... | .................... | .................... | .................... | .................... |
| J | 2774 hCoV-19/England/ALDP-94FB38/2020\|EPI_ISL_555182\|20 | .................... | .................... | .................... | .................... | .................... | .................... |
| J | 2775 hCoV-19/England/ALDP-94FE4E/2020\|EPI_ISL_555183\|20 | .................... | .................... | .................... | .................... | .................... | .................... |
| J | 2776 hCoV-19/England/ALDP-9503C3/2020\|EPI_ISL_555184\|20 | .................... | .................... | .................... | .................... | .................... | .................... |

J 2777 hCoV-19/England/ALDP-94FD23/2020|EPI_ISL_555185|20 ···················· ···················· ····················· ···················· ···················· ····················
J 2778 hCoV-19/England/ALDP-94D9A9/2020|EPI_ISL_555187|20 ···················· ···················· ····················· ···················· ···················· ····················
J 2779 hCoV-19/England/ALDP-94FB0B/2020|EPI_ISL_555188|20 ···················· ···················· ····················· ···················· ···················· ····················
J 2780 hCoV-19/England/ALDP-94E331/2020|EPI_ISL_555189|20 ···················· ···················· ····················· ···················· ···················· ····················
J 2781 hCoV-19/Wales/ALDP-9503D2/2020|EPI_ISL_555191|2020 ···················· ···················· ····················· ···················· ···················· ····················
J 2782 hCoV-19/England/ALDP-94D8F7/2020|EPI_ISL_555192|20 ···················· ···················· ····················· ···················· ···················· ····················
J 2783 hCoV-19/England/ALDP-950053/2020|EPI_ISL_555193|20 ···················· ···················· ····················· ···················· ···················· ····················
J 2784 hCoV-19/England/ALDP-94FCF9/2020|EPI_ISL_555194|20 ···················· ···················· ····················· ···················· ···················· ····················
J 2785 hCoV-19/England/ALDP-94FA4A/2020|EPI_ISL_555195|20 ···················· ···················· ····················· ···················· ···················· ····················
J 2786 hCoV-19/England/ALDP-94D903/2020|EPI_ISL_555196|20 ···················· ···················· ····················· ···················· ···················· ····················
J 2787 hCoV-19/England/ALDP-94E313/2020|EPI_ISL_555197|20 ···················· ···················· ····················· ···················· ···················· ····················
J 2788 hCoV-19/England/ALDP-94FC08/2020|EPI_ISL_555198|20 ···················· ···················· ····················· ···················· ···················· ····················
J 2789 hCoV-19/England/ALDP-9504A2/2020|EPI_ISL_555199|20 ···················· ···················· ····················· ···················· ···················· ····················
J 2790 hCoV-19/England/ALDP-94A441/2020|EPI_ISL_556470|20 ···················· ···················· ····················· ···················· ···················· ····················
J 2791 hCoV-19/England/CAMC-94B03D/2020|EPI_ISL_556471|20 ···················· ···················· ···········T········· ···················· ···················· ····················
J 2792 hCoV-19/England/ALDP-94CB68/2020|EPI_ISL_556473|20 ···················· ···················· ····················· ···················· ···················· ····················
J 2793 hCoV-19/England/ALDP-94EC45/2020|EPI_ISL_556474|20 ···················· ···················· ····················· ···················· ···················· ····················
J 2794 hCoV-19/England/ALDP-94EE21/2020|EPI_ISL_556475|20 ···················· ·A·················· ····················· ···················· ···················· ····················
J 2795 hCoV-19/England/ALDP-94D0C2/2020|EPI_ISL_556476|20 ···················· ···················· ····················· ···················· ···················· ····················
J 2796 hCoV-19/England/ALDP-94999E/2020|EPI_ISL_556477|20 ···················· ···················· ····················· ···················· ···················· ····················
J 2797 hCoV-19/England/ALDP-94ED42/2020|EPI_ISL_556478|20 ···················· ···················· ····················· ···················· ···················· ····················
J 2798 hCoV-19/England/ALDP-94D44E/2020|EPI_ISL_556479|20 ···················· ···················· ····················· ···················· ···················· ····················
J 2799 hCoV-19/England/ALDP-94AEBC/2020|EPI_ISL_556480|20 ···················· ···················· ····················· ···················· ···················· ····················
J 2800 hCoV-19/England/ALDP-94F190/2020|EPI_ISL_556481|20 ···················· ···················· ····················· ···················· ···················· ····················
J 2801 hCoV-19/England/CAMC-94B6D1/2020|EPI_ISL_556482|20 ···················· ···················· ····················· ···················· ···················· ····················
J 2802 hCoV-19/England/ALDP-949C4A/2020|EPI_ISL_556483|20 ···················· ···················· ····················· ···················· ···················· ····················
J 2803 hCoV-19/England/ALDP-94EBB1/2020|EPI_ISL_556484|20 ···················· ···················· ····················· ···················· ···················· ····················
J 2804 hCoV-19/England/ALDP-94D0B3/2020|EPI_ISL_556485|20 ···················· ···················· ····················· ···················· ···················· ····················
J 2805 hCoV-19/England/ALDP-94A380/2020|EPI_ISL_556486|20 ···················· ···················· ····················· ···················· ···················· ····················
J 2806 hCoV-19/England/ALDP-949A13/2020|EPI_ISL_556487|20 ···················· ···················· ····················· ···················· ···················· ····················
J 2807 hCoV-19/England/CAMC-94B5D4/2020|EPI_ISL_556488|20 ···················· ···················· ····················· ···················· ···················· ····················
J 2808 hCoV-19/England/ALDP-94F330/2020|EPI_ISL_556489|20 ···················· ···················· ····················· ···················· ···················· ····················
J 2809 hCoV-19/England/ALDP-94ABB5/2020|EPI_ISL_556490|20 ···················· ···················· ····················· ···················· ···················· ····················
J 2810 hCoV-19/England/ALDP-94B510/2020|EPI_ISL_556491|20 ···················· ···················· ····················· ···················· ···················· ····················
J 2811 hCoV-19/England/ALDP-949873/2020|EPI_ISL_556492|20 ···················· ···················· ····················· ···················· ···················· ····················
J 2812 hCoV-19/England/ALDP-94A344/2020|EPI_ISL_556493|20 ···················· ···················· ····················· ···················· ···················· ····················
J 2813 hCoV-19/England/CAMC-94908A/2020|EPI_ISL_556494|20 ···················· ···················· ····················· ···················· ···················· ····················
J 2814 hCoV-19/England/ALDP-94EE9A/2020|EPI_ISL_556496|20 ···················· ···················· ····················· ···················· ···················· ····················
J 2815 hCoV-19/England/ALDP-94D38D/2020|EPI_ISL_556498|20 ···················· ·······G············ ····················· ···················· ···················· ····················
J 2816 hCoV-19/England/ALDP-94D314/2020|EPI_ISL_556499|20 ···················· ···················· ····················· ···················· ···················· ····················
J 2817 hCoV-19/England/CAMB-1B5D8A/2020|EPI_ISL_549389|20 ···················· ·A·················· ····················· ···················· ···················· ···A················
J 2818 hCoV-19/England/CAMB-1B5DA8/2020|EPI_ISL_549390|20 ···················· ···················· ····················· ···················· ···················· ····················
J 2819 hCoV-19/England/CAMB-1B5DC6/2020|EPI_ISL_549391|20 ···················· ···················· ····················· ···················· ······T············· ····················
J 2820 hCoV-19/England/CAMB-1B5DD5/2020|EPI_ISL_549392|20 ···················· ······C············· ····················· ···················· ···················· ····················
J 2821 hCoV-19/England/CAMB-1B5DE4/2020|EPI_ISL_549393|20 ···················· ······C············· ····················· ···················· ···················· ····················
J 2822 hCoV-19/England/CAMB-1B5E1E/2020|EPI_ISL_549394|20 ···················· ···················· ····················· ···················· ······T············· ····················
J 2823 hCoV-19/England/CAMB-1B5EE1/2020|EPI_ISL_549395|20 ···················· ···················· ····················· ···················· ···················· ····················
J 2824 hCoV-19/England/CAMC-9B1E38/2020|EPI_ISL_549396|20 ···················· ···················· ····················· ···················· ···················· ····················
J 2825 hCoV-19/England/CAMC-9B1E65/2020|EPI_ISL_549397|20 ···················· ···················· ····················· ···················· ···················· ····················
J 2826 hCoV-19/England/CAMC-9B1E92/2020|EPI_ISL_549398|20 ···················· ···················· ····················· ···················· ···················· ····················
J 2827 hCoV-19/England/CAMC-9B1EA1/2020|EPI_ISL_549399|20 ···················· ···················· ····················· ···················· ···················· ····················
J 2828 hCoV-19/England/MILK-99A03F/2020|EPI_ISL_550365|20 ···················· ···················· ····················· ···················· ···················· ····················
J 2829 hCoV-19/England/MILK-999EAF/2020|EPI_ISL_550366|20 ···················· ···················· ····················· ···················· ···················· ····················
J 2830 hCoV-19/England/MILK-99A530/2020|EPI_ISL_550369|20 ···················· ···················· ····················· ···················· ···················· ····················
J 2831 hCoV-19/England/MILK-99A99E/2020|EPI_ISL_550372|20 ···················· ···················· ····················· ···················· ···················· ····················
J 2832 hCoV-19/Wales/MILK-999CD2/2020|EPI_ISL_550373|2020 ···················· ···················· ····················· ···················· ···················· ····················
J 2833 hCoV-19/England/MILK-99A178/2020|EPI_ISL_550376|20 ···················· ···················· ····················· ···················· ···················· ····················
J 2834 hCoV-19/England/MILK-99B55D/2020|EPI_ISL_550377|20 ···················· ···················· ····················· ···················· ···················· ····················
J 2835 hCoV-19/England/MILK-99A7EF/2020|EPI_ISL_550378|20 ···················· ···················· ····················· ···················· ···················· ····················
J 2836 hCoV-19/England/MILK-99A020/2020|EPI_ISL_550379|20 ···················· ···················· ····················· ···················· ···················· ····················
J 2837 hCoV-19/England/MILK-999D84/2020|EPI_ISL_550380|20 ···················· ···················· ····················· ···················· ···················· ····················
J 2838 hCoV-19/England/MILK-99A5C7/2020|EPI_ISL_550381|20 ···················· ···················· ····················· ···················· ···················· ····················
J 2839 hCoV-19/England/MILK-99A04E/2020|EPI_ISL_550382|20 ···················· ···················· ····················· ···················· ···················· ····················
J 2840 hCoV-19/England/MILK-99A61F/2020|EPI_ISL_550383|20 ···················· ···················· ····················· ···················· ···················· ····················
J 2841 hCoV-19/England/MILK-999FF7/2020|EPI_ISL_550385|20 ···················· ···················· ····················· ···················· ···················· ····················
J 2842 hCoV-19/England/MILK-99A9AD/2020|EPI_ISL_550386|20 ···················· ···················· ····················· ···················· ···················· ····················
J 2843 hCoV-19/England/MILK-99A460/2020|EPI_ISL_550387|20 ···················· ···················· ····················· ···················· ···················· ····················
J 2844 hCoV-19/England/MILK-99A65B/2020|EPI_ISL_550388|20 ···················· ···················· ····················· ···················· ···················· ····················
J 2845 hCoV-19/England/MILK-99A785/2020|EPI_ISL_550389|20 ···················· ·A·······A·········· ····················· ···················· ···················· ····················
J 2846 hCoV-19/England/MILK-99A266/2020|EPI_ISL_550390|20 ···················· ···················· ····················· ···················· ···················· ····················
J 2847 hCoV-19/England/MILK-999D0C/2020|EPI_ISL_550391|20 ···················· ···················· ····················· ···················· ···················· ····················
J 2848 hCoV-19/England/MILK-99A1C3/2020|EPI_ISL_550392|20 ···················· ···················· ····················· ···················· ···················· ····················
J 2849 hCoV-19/England/MILK-99A9F8/2020|EPI_ISL_550394|20 ···················· ···················· ····················· ···················· ···················· ····················
J 2850 hCoV-19/Wales/MILK-999CF0/2020|EPI_ISL_550396|2020 ···················· ···················· ····················· ···················· ···················· ····················
J 2851 hCoV-19/Wales/MILK-99A697/2020|EPI_ISL_550397|2020 ···················· ···················· ····················· ···················· ···················· ····················
J 2852 hCoV-19/England/MILK-999ECD/2020|EPI_ISL_550398|20 ···················· ···················· ····················· ···················· ···················· ····················
J 2853 hCoV-19/England/MILK-99A5A9/2020|EPI_ISL_550399|20 ···················· ···················· ····················· ···················· ···················· ····················
J 2854 hCoV-19/England/MILK-995C5E/2020|EPI_ISL_551600|20 ···················· ···················· ····················· ···················· ···················· ····················
J 2855 hCoV-19/England/MILK-99433C/2020|EPI_ISL_551601|20 ···················· ·A·················· ····················· ···················· ···················· ···A················
J 2856 hCoV-19/Wales/MILK-9955DB/2020|EPI_ISL_551603|2020 ···················· ···················· ····················· ···················· ···················· ····················

| | | | | | | | | |
|---|---|---|---|---|---|---|---|---|
| J | 2857 | hCoV-19/England/MILK-995C4F/2020\|EPI_ISL_551604\|20 | ···················· | ···················· | ····················· | ···················· | ···················· | ···················· |
| J | 2858 | hCoV-19/England/MILK-995B24/2020\|EPI_ISL_551605\|20 | ···················· | ···················· | ····················· | ···················· | ···················· | ···················· |
| J | 2859 | hCoV-19/England/MILK-993E87/2020\|EPI_ISL_551606\|20 | ···················· | ···················· | ····················· | ···················· | ···················· | ···················· |
| J | 2860 | hCoV-19/England/MILK-995ADC/2020\|EPI_ISL_551607\|20 | ···················· | ···················· | ····················· | ···················· | ···················· | ···················· |
| J | 2861 | hCoV-19/England/MILK-994396/2020\|EPI_ISL_551611\|20 | ···················· | ···················· | ····················· | ···················· | ···················· | ···················· |
| J | 2862 | hCoV-19/England/MILK-9942B7/2020\|EPI_ISL_551612\|20 | ···················· | ···················· | ····················· | ···················· | ···················· | ···················· |
| J | 2863 | hCoV-19/England/MILK-993FDF/2020\|EPI_ISL_551613\|20 | ···················· | ···················· | ····················· | ···················· | ···················· | ···················· |
| J | 2864 | hCoV-19/England/MILK-995508/2020\|EPI_ISL_551614\|20 | ···················· | ···················· | ····················· | ···················· | ···················· | ···················· |
| J | 2865 | hCoV-19/England/MILK-993E3C/2020\|EPI_ISL_551615\|20 | ···················· | ···················· | ····················· | ···················· | ···················· | ···················· |
| J | 2866 | hCoV-19/England/MILK-99424E/2020\|EPI_ISL_551616\|20 | ···················· | ···················· | ····················· | ···················· | ···················· | ···················· |
| J | 2867 | hCoV-19/England/MILK-9959A2/2020\|EPI_ISL_551617\|20 | ···················· | ···················· | ····················· | ···················· | ···················· | ···················· |
| J | 2868 | hCoV-19/England/MILK-99426C/2020\|EPI_ISL_551618\|20 | ···················· | ···················· | ····················· | ···················· | ···················· | ···················· |
| J | 2869 | hCoV-19/Wales/MILK-995C03/2020\|EPI_ISL_551627\|2020 | ···················· | ···················· | ····················· | ···················· | ···················· | ···················· |
| J | 2870 | hCoV-19/Wales/MILK-995BF7/2020\|EPI_ISL_551696\|2020 | ···················· | ···················· | ····················· | ···················· | ···················· | ···················· |
| J | 2871 | hCoV-19/England/MILK-995614/2020\|EPI_ISL_551697\|20 | ···················· | ···················· | ····················· | ···················· | ···················· | ···················· |
| J | 2872 | hCoV-19/England/MILK-9941AB/2020\|EPI_ISL_551699\|20 | ···················· | ···················· | ····················· | ···················· | ···················· | ···················· |
| J | 2873 | hCoV-19/England/ALDP-924191/2020\|EPI_ISL_552912\|20 | ···················· | ···················· | ····················· | ···················· | ···················· | ···················· |
| J | 2874 | hCoV-19/England/ALDP-93036F/2020\|EPI_ISL_552913\|20 | ···················· | ···················· | ····················· | ···················· | ···················· | ···················· |
| J | 2875 | hCoV-19/England/ALDP-936E2C/2020\|EPI_ISL_552921\|20 | ···················· | ···················· | ····················· | ···················· | ···················· | ···················· |
| J | 2876 | hCoV-19/England/ALDP-928C6F/2020\|EPI_ISL_552922\|20 | ···················· | ···················· | ····················· | ···················· | ···················· | ···················· |
| J | 2877 | hCoV-19/England/ALDP-920D28/2020\|EPI_ISL_552923\|20 | ···················· | ·A·················· | ····················· | ···················· | ···················· | ···················· |
| J | 2878 | hCoV-19/England/ALDP-935ACF/2020\|EPI_ISL_552925\|20 | ···················· | ···················· | ····················· | ···················· | ···················· | ···················· |
| J | 2879 | hCoV-19/England/ALDP-931270/2020\|EPI_ISL_552926\|20 | ···················· | ···················· | ····················· | ···················· | ···················· | ···················· |
| J | 2880 | hCoV-19/England/ALDP-92B570/2020\|EPI_ISL_552927\|20 | ···················· | ···················· | ····················· | ···················· | ···················· | ···················· |
| J | 2881 | hCoV-19/England/ALDP-939CC5/2020\|EPI_ISL_552928\|20 | ···················· | ···················· | ····················· | ···················· | ···················· | ···················· |
| J | 2882 | hCoV-19/England/ALDP-92677F/2020\|EPI_ISL_552929\|20 | ···················· | ···················· | ····················· | ···················· | ···················· | ···················· |
| J | 2883 | hCoV-19/England/ALDP-929220/2020\|EPI_ISL_552930\|20 | ···················· | ···················· | ····················· | ···················· | ···················· | ···················· |
| J | 2884 | hCoV-19/England/ALDP-920E70/2020\|EPI_ISL_552931\|20 | ···················· | ·A·················· | ····················· | ···················· | ···················· | ···················· |
| J | 2885 | hCoV-19/England/ALDP-9348B7/2020\|EPI_ISL_552933\|20 | ···················· | ···················· | ····················· | ···················· | ···················· | ···················· |
| J | 2886 | hCoV-19/England/ALDP-936BE9/2020\|EPI_ISL_552934\|20 | ···················· | ···················· | ····················· | ···················· | ···················· | ···················· |
| J | 2887 | hCoV-19/England/ALDP-9256A0/2020\|EPI_ISL_552935\|20 | ···················· | ···················· | ····················· | ···················· | ···················· | ···················· |
| J | 2888 | hCoV-19/England/MILK-93D3BD/2020\|EPI_ISL_552936\|20 | ···················· | ···················· | ····················· | ···················· | ···················· | ···················· |
| J | 2889 | hCoV-19/England/ALDP-93578C/2020\|EPI_ISL_552937\|20 | ···················· | ·A·················· | ····················· | ···················· | ···················· | ···················· |
| J | 2890 | hCoV-19/England/MILK-93D678/2020\|EPI_ISL_552938\|20 | ···················· | ···················· | ····················· | ···················· | ···················· | ···················· |
| J | 2891 | hCoV-19/England/MILK-93D10E/2020\|EPI_ISL_552939\|20 | ···················· | ···················· | ····················· | ···················· | ···················· | ···················· |
| J | 2893 | hCoV-19/England/ALDP-92841C/2020\|EPI_ISL_552941\|20 | ···················· | ···················· | ····················· | ···················· | ···················· | ···················· |
| J | 2894 | hCoV-19/England/ALDP-9032A4/2020\|EPI_ISL_552942\|20 | ···················· | ···················· | ····················· | ···················· | ···················· | ···················· |
| J | 2895 | hCoV-19/England/ALDP-9240D0/2020\|EPI_ISL_552944\|20 | ···················· | ···················· | ····················· | ···················· | ···················· | ···················· |
| J | 2896 | hCoV-19/England/ALDP-9336BE/2020\|EPI_ISL_552945\|20 | ···················· | ···················· | ····················· | ···················· | ···················· | ···················· |
| J | 2897 | hCoV-19/England/ALDP-920C67/2020\|EPI_ISL_552946\|20 | ···················· | ···················· | ····················· | ···················· | ···················· | ···················· |
| J | 2898 | hCoV-19/England/MILK-93BB8A/2020\|EPI_ISL_552947\|20 | ···················· | ···················· | ····················· | ···················· | ···················· | ···················· |
| J | 2899 | hCoV-19/England/MILK-93D8FA/2020\|EPI_ISL_552948\|20 | ···················· | ···················· | ····················· | ···················· | ···················· | ···················· |
| J | 2900 | hCoV-19/England/MILK-93D3EA/2020\|EPI_ISL_552949\|20 | ···················· | ···················· | ····················· | ···················· | ···················· | ···················· |
| J | 2901 | hCoV-19/England/ALDP-95AA6A/2020\|EPI_ISL_555100\|20 | ···················· | ···················· | ····················· | ···················· | ···················· | ···················· |
| J | 2902 | hCoV-19/England/ALDP-95C0EE/2020\|EPI_ISL_555101\|20 | ···················· | ···················· | ····················· | ···················· | ···················· | ···················· |
| J | 2903 | hCoV-19/England/ALDP-95C5FE/2020\|EPI_ISL_555102\|20 | ···················· | ···················· | ····················· | ···················· | ···················· | ···················· |
| J | 2904 | hCoV-19/Wales/ALDP-94D7AF/2020\|EPI_ISL_555103\|2020 | ···················· | ···················· | ····················· | ···················· | ···················· | ···················· |
| J | 2905 | hCoV-19/England/ALDP-94E4A7/2020\|EPI_ISL_555104\|20 | ···················· | ···················· | ····················· | ···················· | ···················· | ···················· |
| J | 2906 | hCoV-19/England/ALDP-94D4F3/2020\|EPI_ISL_555106\|20 | ···················· | ···················· | ····················· | ···················· | ···················· | ···················· |
| J | 2907 | hCoV-19/England/ALDP-95C145/2020\|EPI_ISL_555107\|20 | ···················· | ···················· | ····················· | ···················· | ···················· | ···················· |
| J | 2908 | hCoV-19/England/ALDP-95C136/2020\|EPI_ISL_555108\|20 | ···················· | ···················· | ····················· | ···················· | ···················· | ···················· |
| J | 2909 | hCoV-19/England/ALDP-95C619/2020\|EPI_ISL_555110\|20 | ···················· | ···················· | ····················· | ···················· | ···················· | ···················· |
| J | 2910 | hCoV-19/England/ALDP-959D26/2020\|EPI_ISL_555111\|20 | ···················· | ···················· | ····················· | ···················· | ···················· | ···················· |
| J | 2911 | hCoV-19/England/ALDP-95C831/2020\|EPI_ISL_555112\|20 | ···················· | ···················· | ····················· | ···················· | ···················· | ···················· |
| J | 2912 | hCoV-19/England/ALDP-95CAC2/2020\|EPI_ISL_555113\|20 | ···················· | ···················· | ····················· | ···················· | ···················· | ···················· |
| J | 2913 | hCoV-19/England/ALDP-95A217/2020\|EPI_ISL_555114\|20 | ···················· | ···················· | ····················· | ···················· | ···················· | ···················· |
| J | 2914 | hCoV-19/England/ALDP-94D8AC/2020\|EPI_ISL_555115\|20 | ···················· | ···················· | ····················· | ···················· | ···················· | ···················· |
| J | 2915 | hCoV-19/England/ALDP-95C242/2020\|EPI_ISL_555116\|20 | ···················· | ···················· | ····················· | ···················· | ···················· | ···················· |
| J | 2916 | hCoV-19/England/ALDP-95D01A/2020\|EPI_ISL_555117\|20 | ···················· | ···················· | ····················· | ···················· | ···················· | ···················· |
| J | 2917 | hCoV-19/England/ALDP-95A235/2020\|EPI_ISL_555118\|20 | ···················· | ···················· | ····················· | ···················· | ···················· | ···················· |
| J | 2918 | hCoV-19/England/ALDP-95CCCC/2020\|EPI_ISL_555119\|20 | ···················· | ···················· | ····················· | ···················· | ···················· | ···················· |
| J | 2919 | hCoV-19/England/ALDP-95C1AF/2020\|EPI_ISL_555120\|20 | ···················· | ···················· | ····················· | ···················· | ···················· | ···················· |
| J | 2920 | hCoV-19/England/ALDP-95A772/2020\|EPI_ISL_555121\|20 | ···················· | ···················· | ····················· | ···················· | ···················· | ···················· |
| J | 2921 | hCoV-19/England/ALDP-94E902/2020\|EPI_ISL_555122\|20 | ···················· | ···················· | ····················· | ···················· | ···················· | ···················· |
| J | 2922 | hCoV-19/England/ALDP-95C8E6/2020\|EPI_ISL_555123\|20 | ···················· | ···················· | ····················· | ···················· | ···················· | ···················· |
| J | 2923 | hCoV-19/England/ALDP-95CD32/2020\|EPI_ISL_555124\|20 | ···················· | ···················· | ····················· | ···················· | ···················· | ···················· |
| J | 2924 | hCoV-19/England/ALDP-95D00B/2020\|EPI_ISL_555125\|20 | ···················· | ···················· | ····················· | ···················· | ···················· | ···················· |
| J | 2925 | hCoV-19/England/ALDP-95CDF6/2020\|EPI_ISL_555126\|20 | ···················· | ···················· | ····················· | ···················· | ···················· | ···················· |
| J | 2926 | hCoV-19/England/ALDP-950518/2020\|EPI_ISL_555130\|20 | ···················· | ···················· | ····················· | ···················· | ···················· | ···················· |
| J | 2927 | hCoV-19/England/ALDP-94F9E3/2020\|EPI_ISL_555131\|20 | ···················· | ···················· | ····················· | ···················· | ···················· | ···················· |
| J | 2928 | hCoV-19/England/ALDP-94F87D/2020\|EPI_ISL_555132\|20 | ···················· | ···················· | ····················· | ···················· | ···················· | ···················· |
| J | 2929 | hCoV-19/England/ALDP-94FEB7/2020\|EPI_ISL_555133\|20 | ···················· | ···················· | ····················· | ···················· | ···················· | ···················· |
| J | 2930 | hCoV-19/England/ALDP-950220/2020\|EPI_ISL_555134\|20 | ···················· | ·A·················· | ····················· | ···················· | ···················· | ···················· |
| J | 2931 | hCoV-19/England/ALDP-94FA0E/2020\|EPI_ISL_555135\|20 | ···················· | ···················· | ····················· | ···················· | ···················· | ···················· |
| J | 2932 | hCoV-19/England/ALDP-950624/2020\|EPI_ISL_555136\|20 | ···················· | ···················· | ····················· | ···················· | ···················· | ···················· |
| J | 2933 | hCoV-19/England/ALDP-9506D9/2020\|EPI_ISL_555137\|20 | ···················· | ···················· | ····················· | ···················· | ···················· | ···················· |
| J | 2934 | hCoV-19/England/ALDP-94A56C/2020\|EPI_ISL_556400\|20 | ···················· | ···················· | ····················· | ···················· | ···················· | ···················· |
| J | 2935 | hCoV-19/England/ALDP-94D36F/2020\|EPI_ISL_556402\|20 | ···················· | ···················· | ····················· | ···················· | ···················· | ···················· |
| J | 2936 | hCoV-19/England/ALDP-94EC72/2020\|EPI_ISL_556405\|20 | ···················· | ···················· | ····················· | ···················· | ···················· | ···················· |
| J | 2937 | hCoV-19/England/ALDP-94ABE2/2020\|EPI_ISL_556406\|20 | ···················· | ···················· | ····················· | ···················· | ···················· | ···················· |

| | | | | | | | | |
|---|---|---|---|---|---|---|---|---|
| J | 2938 | hCoV-19/England/ALDP-94F066/2020\|EPI_ISL_556407\|20 | ···················· | ···················· | ···················· | ···················· | ···················· | ···················· |
| J | 2939 | hCoV-19/England/ALDP-94A405/2020\|EPI_ISL_556408\|20 | ···················· | ···················· | ···················· | ···················· | ···················· | ···················· |
| J | 2940 | hCoV-19/England/ALDP-94F1CD/2020\|EPI_ISL_556410\|20 | ···················· | ···················· | ···················· | ···················· | ···················· | ···················· |
| J | 2941 | hCoV-19/England/ALDP-94ED8E/2020\|EPI_ISL_556412\|20 | ···················· | ···················· | ···················· | ···················· | ···················· | ···················· |
| J | 2942 | hCoV-19/England/ALDP-94EE5E/2020\|EPI_ISL_556413\|20 | ···················· | ···················· | ···················· | ···················· | ···················· | ···················· |
| J | 2943 | hCoV-19/England/ALDP-9498DD/2020\|EPI_ISL_556414\|20 | ···················· | ···················· | ···················· | ···················· | ···················· | ···················· |
| J | 2944 | hCoV-19/England/ALDP-94B8F9/2020\|EPI_ISL_556415\|20 | ···················· | ···················· | ···················· | ···················· | ···················· | ···················· |
| J | 2945 | hCoV-19/England/ALDP-94D226/2020\|EPI_ISL_556416\|20 | ···················· | ···················· | ···················· | ···················· | ···················· | ···················· |
| J | 2946 | hCoV-19/England/ALDP-94B255/2020\|EPI_ISL_556417\|20 | ···················· | ···················· | ···················· | ···················· | ···················· | ···················· |
| J | 2947 | hCoV-19/England/ALDP-94B5B6/2020\|EPI_ISL_556418\|20 | ···················· | ···················· | ···················· | ···················· | ···················· | ···················· |
| J | 2948 | hCoV-19/England/ALDP-94A7EE/2020\|EPI_ISL_556419\|20 | ···················· | ···················· | ···················· | ···················· | ···················· | ···················· |
| J | 2949 | hCoV-19/England/ALDP-94D341/2020\|EPI_ISL_556420\|20 | ···················· | ···················· | ···················· | ···················· | ···················· | ···················· |
| J | 2950 | hCoV-19/England/ALDP-94A450/2020\|EPI_ISL_556421\|20 | ···················· | ···················· | ···················· | ···················· | ···················· | ···················· |
| J | 2951 | hCoV-19/England/ALDP-94D1ED/2020\|EPI_ISL_556422\|20 | ···················· | ···················· | ···················· | ···················· | ···················· | ···················· |
| J | 2952 | hCoV-19/England/ALDP-94AA8B/2020\|EPI_ISL_556424\|20 | ···················· | ···················· | ···················· | ···················· | ···················· | ···················· |
| J | 2953 | hCoV-19/England/CAMC-94B1FE/2020\|EPI_ISL_556425\|20 | ···················· | ···················· | ···················· | ···················· | ···················· | ···················· |
| J | 2954 | hCoV-19/England/ALDP-949C3B/2020\|EPI_ISL_556426\|20 | ···················· | ···················· | ···················· | ···················· | ···················· | ···················· |
| J | 2955 | hCoV-19/England/ALDP-94B5C5/2020\|EPI_ISL_556427\|20 | ···················· | ·A·················· | ···················· | ···················· | ···················· | ···················· |
| J | 2956 | hCoV-19/England/ALDP-94F1DC/2020\|EPI_ISL_556429\|20 | ···················· | ···················· | ···················· | ···················· | ···················· | ···················· |
| J | 2957 | hCoV-19/England/ALDP-94EFE2/2020\|EPI_ISL_556430\|20 | ···················· | ···················· | ···················· | ···················· | ···················· | ···················· |
| J | 2958 | hCoV-19/England/ALDP-94A098/2020\|EPI_ISL_556431\|20 | ···················· | ···················· | ···················· | ···················· | ···················· | ···················· |
| J | 2959 | hCoV-19/England/CAMC-94A7A2/2020\|EPI_ISL_556434\|20 | ···················· | ···················· | ···················· | ···················· | ······T············· | ···················· |
| J | 2960 | hCoV-19/England/ALDP-94ED60/2020\|EPI_ISL_556436\|20 | ···················· | ···················· | ···················· | ···················· | ···················· | ···················· |
| J | 2961 | hCoV-19/England/ALDP-94AF04/2020\|EPI_ISL_556437\|20 | ···················· | ···················· | ···················· | ···················· | ···················· | ···················· |
| J | 2962 | hCoV-19/England/ALDP-94F46A/2020\|EPI_ISL_556438\|20 | ···················· | ···················· | ···················· | ···················· | ···················· | ···················· |
| J | 2963 | hCoV-19/England/ALDP-94EE8B/2020\|EPI_ISL_556440\|20 | ···················· | ···················· | ···················· | ···················· | ···················· | ···················· |
| J | 2964 | hCoV-19/England/ALDP-94D39C/2020\|EPI_ISL_556441\|20 | ···················· | ···················· | ···················· | ···················· | ···················· | ···················· |
| J | 2965 | hCoV-19/England/ALDP-94D0A4/2020\|EPI_ISL_556443\|20 | ···················· | ···················· | ···················· | ···················· | ···················· | ···················· |
| J | 2966 | hCoV-19/England/ALDP-94F1FA/2020\|EPI_ISL_556444\|20 | ···················· | ···················· | ···················· | ···················· | ···················· | ···················· |
| J | 2967 | hCoV-19/England/CAMC-94B853/2020\|EPI_ISL_556446\|20 | ···················· | ···················· | ···················· | ···················· | ···················· | ···················· |
| J | 2968 | hCoV-19/England/ALDP-94A4BA/2020\|EPI_ISL_556448\|20 | ···················· | ···················· | ···················· | ···················· | ···················· | ···················· |
| J | 2969 | hCoV-19/England/ALDP-94F0C0/2020\|EPI_ISL_556449\|20 | ···················· | ···················· | ···················· | ···················· | ···················· | ···················· |
| J | 2970 | hCoV-19/England/ALDP-94A6D2/2020\|EPI_ISL_556450\|20 | ···················· | ···················· | ···················· | ···················· | ···················· | ···················· |
| J | 2971 | hCoV-19/England/ALDP-94EF88/2020\|EPI_ISL_556451\|20 | ···················· | ···················· | ···················· | ···················· | ···················· | ···················· |
| J | 2972 | hCoV-19/England/ALDP-94AF6E/2020\|EPI_ISL_556452\|20 | ···················· | ···················· | ···················· | ···················· | ···················· | ···················· |
| J | 2973 | hCoV-19/England/ALDP-94AEE9/2020\|EPI_ISL_556454\|20 | ···················· | ···················· | ···················· | ···················· | ···················· | ···················· |
| J | 2974 | hCoV-19/England/ALDP-94D271/2020\|EPI_ISL_556455\|20 | ···················· | ···················· | ···················· | ···················· | ···················· | ···················· |
| J | 2975 | hCoV-19/England/ALDP-94ABC4/2020\|EPI_ISL_556456\|20 | ···················· | ···················· | ···················· | ···················· | ···················· | ···················· |
| J | 2976 | hCoV-19/England/ALDP-94AEF8/2020\|EPI_ISL_556457\|20 | ···················· | ···················· | ···················· | ···················· | ···················· | ···················· |
| J | 2977 | hCoV-19/England/CAMC-94AAD6/2020\|EPI_ISL_556459\|20 | ···················· | ·A·················· | ···················· | ···················· | ···················· | ···A················ |
| J | 2978 | hCoV-19/England/ALDP-94F1EB/2020\|EPI_ISL_556460\|20 | ···················· | ···················· | ···················· | ···················· | ···················· | ···················· |
| J | 2979 | hCoV-19/England/ALDP-949D38/2020\|EPI_ISL_556461\|20 | ···················· | ···················· | ···················· | ···················· | ···················· | ···················· |
| J | 2980 | hCoV-19/England/ALDP-94B8DB/2020\|EPI_ISL_556462\|20 | ···················· | ···················· | ···················· | ···················· | ···················· | ···················· |
| J | 2981 | hCoV-19/England/ALDP-94EE7C/2020\|EPI_ISL_556463\|20 | ···················· | ···················· | ···················· | ···················· | ···················· | ···················· |
| J | 2982 | hCoV-19/England/ALDP-94EB93/2020\|EPI_ISL_556464\|20 | ···················· | ·A·················· | ···················· | ···················· | ···················· | ···················· |
| J | 2983 | hCoV-19/England/ALDP-94B1C1/2020\|EPI_ISL_556465\|20 | ···················· | ···················· | ···················· | ···················· | ···················· | ···················· |
| J | 2984 | hCoV-19/England/ALDP-94D3E7/2020\|EPI_ISL_556467\|20 | ···················· | ···················· | ···················· | ···················· | ···················· | ···················· |
| J | 2985 | hCoV-19/England/ALDP-94A03E/2020\|EPI_ISL_556468\|20 | ···················· | ···················· | ···················· | ···················· | ···················· | ···················· |
| J | 2986 | hCoV-19/England/ALDP-94A836/2020\|EPI_ISL_556469\|20 | ···················· | ···················· | ···················· | ···················· | ···················· | ···················· |
| J | 2987 | hCoV-19/Wales/ALDP-94F039/2020\|EPI_ISL_556501\|2020 | ···················· | ···················· | ···················· | ···················· | ···················· | ···················· |
| J | 2988 | hCoV-19/England/ALDP-94EEB8/2020\|EPI_ISL_556502\|20 | ···················· | ···················· | ···················· | ···················· | ···················· | ···················· |
| J | 2989 | hCoV-19/England/ALDP-6FF8BB/2020\|EPI_ISL_557714\|20 | ···················· | ···················· | ···················· | ···················· | ···················· | ···················· |
| J | 2990 | hCoV-19/England/ALDP-71C50B/2020\|EPI_ISL_557715\|20 | ···················· | ···················· | ···················· | ···················· | ···················· | ···················· |
| J | 2991 | hCoV-19/England/ALDP-7000BE/2020\|EPI_ISL_557717\|20 | ···················· | ···················· | ···················· | ···················· | ···················· | ···················· |
| J | 2992 | hCoV-19/England/ALDP-704BE6/2020\|EPI_ISL_557718\|20 | ···················· | ···················· | ···················· | ···················· | ···················· | ···················· |
| J | 2993 | hCoV-19/England/ALDP-6E82D5/2020\|EPI_ISL_557719\|20 | ···················· | ···················· | ···················· | ···················· | ···················· | ···················· |
| J | 2994 | hCoV-19/England/ALDP-6DE2C3/2020\|EPI_ISL_557720\|20 | ···················· | ···················· | ···················· | ···················· | ···················· | ···················· |
| J | 2995 | hCoV-19/England/ALDP-725AB5/2020\|EPI_ISL_557721\|20 | ···················· | ···················· | ···················· | ···················· | ···················· | ···················· |
| J | 2996 | hCoV-19/England/ALDP-71CBAF/2020\|EPI_ISL_557722\|20 | ···················· | ···················· | ···················· | ···················· | ···················· | ···················· |
| J | 2997 | hCoV-19/England/ALDP-6DF918/2020\|EPI_ISL_557723\|20 | ···················· | ···················· | ···················· | ···················· | ···················· | ···················· |
| J | 2998 | hCoV-19/England/ALDP-71D9C2/2020\|EPI_ISL_557724\|20 | ···················· | ···················· | ···················· | ···················· | ···················· | ···················· |
| J | 2999 | hCoV-19/England/ALDP-71D124/2020\|EPI_ISL_557725\|20 | ···················· | ···················· | ···················· | ···················· | ···················· | ···················· |
| J | 3000 | hCoV-19/England/ALDP-71E0DB/2020\|EPI_ISL_557727\|20 | ···················· | ···················· | ···················· | ···················· | ···················· | ···················· |
| J | 3001 | hCoV-19/England/ALDP-71C4B3/2020\|EPI_ISL_557729\|20 | ···················· | ···················· | ···················· | ···················· | ···················· | ···················· |
| J | 3002 | hCoV-19/England/ALDP-6EAEDF/2020\|EPI_ISL_557731\|20 | ···················· | ···················· | ···················· | ···················· | ···················· | ···················· |
| J | 3003 | hCoV-19/England/ALDP-6E9D00/2020\|EPI_ISL_557733\|20 | ···················· | ···················· | ···················· | ···················· | ···················· | ···················· |
| J | 3004 | hCoV-19/England/ALDP-70A5E2/2020\|EPI_ISL_557734\|20 | ···················· | ···················· | ···················· | ···················· | ···················· | ···················· |
| J | 3005 | hCoV-19/England/ALDP-724FB7/2020\|EPI_ISL_557735\|20 | ···················· | ······A············· | ···················· | ···················· | ···················· | ···················· |
| J | 3006 | hCoV-19/England/ALDP-700C32/2020\|EPI_ISL_557736\|20 | ···················· | ···················· | ···················· | ···················· | ···················· | ···················· |
| J | 3007 | hCoV-19/England/ALDP-70704E/2020\|EPI_ISL_557737\|20 | ···················· | ···················· | ···················· | ···················· | ···················· | ···················· |
| J | 3008 | hCoV-19/England/ALDP-6B5927/2020\|EPI_ISL_557738\|20 | ···················· | ···················· | ···················· | ···················· | ···················· | ···················· |
| J | 3009 | hCoV-19/England/MILK-6395CD/2020\|EPI_ISL_557740\|20 | ···················· | ···················· | ···················· | ···················· | ···················· | ···················· |
| J | 3010 | hCoV-19/England/MILK-6285D0/2020\|EPI_ISL_557742\|20 | ···················· | ···················· | ···················· | ···················· | ···················· | ···················· |
| J | 3011 | hCoV-19/England/ALDP-6BE311/2020\|EPI_ISL_557744\|20 | ···················· | ···················· | ···················· | ···················· | ···················· | ···················· |
| J | 3012 | hCoV-19/England/ALDP-6C176A/2020\|EPI_ISL_557747\|20 | ···················· | ···················· | ···················· | ···················· | ···················· | ···················· |
| J | 3013 | hCoV-19/England/MILK-63AC6D/2020\|EPI_ISL_557748\|20 | ···················· | ···················· | ···················· | ···················· | ···················· | ···················· |
| J | 3014 | hCoV-19/England/ALDP-6BEF68/2020\|EPI_ISL_557749\|20 | ···················· | ···················· | ···················· | ···················· | ···················· | ···················· |
| J | 3015 | hCoV-19/England/ALDP-6BF79C/2020\|EPI_ISL_557751\|20 | ···················· | ···················· | ···················· | ···················· | ···················· | ···················· |
| J | 3016 | hCoV-19/England/MILK-61E045/2020\|EPI_ISL_557753\|20 | ···················· | ···················· | ···················· | ···················· | ···················· | ···················· |
| J | 3017 | hCoV-19/England/ALDP-6ADB68/2020\|EPI_ISL_557754\|20 | ···················· | ···················· | ···················· | ···················· | ···················· | ···················· |

| | | | | | | | | |
|---|---|---|---|---|---|---|---|---|
| J | 3018 | hCoV-19/England/ALDP-6AD342/2020\|EPI_ISL_557755\|20 | ···················· | ···················· | ····················· | ···················· | ···················· | ···················· |
| J | 3019 | hCoV-19/England/ALDP-6AEB0D/2020\|EPI_ISL_557756\|20 | ···················· | ···················· | ····················· | ···················· | ···················· | ···················· |
| J | 3020 | hCoV-19/England/MILK-6210A9/2020\|EPI_ISL_557757\|20 | ···················· | ···················· | ····················· | ···················· | ···················· | ···················· |
| J | 3021 | hCoV-19/England/ALDP-6BC7DB/2020\|EPI_ISL_557759\|20 | ···················· | ···················· | ····················· | ···················· | ···················· | ···················· |
| J | 3022 | hCoV-19/England/ALDP-6BB648/2020\|EPI_ISL_557761\|20 | ···················· | ···················· | ····················· | ···················· | ···················· | ···················· |
| J | 3023 | hCoV-19/England/ALDP-6B715A/2020\|EPI_ISL_557762\|20 | ···················· | ···················· | ····················· | ···················· | ···················· | ···················· |
| J | 3024 | hCoV-19/England/ALDP-6BBC28/2020\|EPI_ISL_557763\|20 | ···················· | ···················· | ····················· | ···················· | ···················· | ···················· |
| J | 3025 | hCoV-19/England/MILK-63931E/2020\|EPI_ISL_557765\|20 | ···················· | ···················· | ····················· | ···················· | ···················· | ···················· |
| J | 3026 | hCoV-19/England/MILK-61E6CB/2020\|EPI_ISL_557766\|20 | ···················· | ···················· | ····················· | ···················· | ···················· | ···················· |
| J | 3027 | hCoV-19/England/ALDP-6AD500/2020\|EPI_ISL_557769\|20 | ···················· | ···················· | ····················· | ···················· | ···················· | ···················· |
| J | 3028 | hCoV-19/England/ALDP-6C1B5F/2020\|EPI_ISL_557770\|20 | ···················· | ···················· | ····················· | ···················· | ···················· | ···················· |
| J | 3029 | hCoV-19/England/ALDP-6AEB2B/2020\|EPI_ISL_557772\|20 | ···················· | ···················· | ····················· | ···················· | ···················· | ···················· |
| J | 3030 | hCoV-19/England/ALDP-6B564E/2020\|EPI_ISL_557773\|20 | ···················· | ···················· | ····················· | ···················· | ···················· | ···················· |
| J | 3031 | hCoV-19/England/ALDP-6BF389/2020\|EPI_ISL_557776\|20 | ···················· | ···················· | ····················· | ···················· | ···················· | ···················· |
| J | 3032 | hCoV-19/England/ALDP-6B0AA1/2020\|EPI_ISL_557779\|20 | ···················· | ···················· | ····················· | ···················· | ···················· | ···················· |
| J | 3033 | hCoV-19/England/ALDP-6AF1DD/2020\|EPI_ISL_557780\|20 | ···················· | ···················· | ····················· | ···················· | ···················· | ···················· |
| J | 3034 | hCoV-19/England/MILK-639D7A/2020\|EPI_ISL_557781\|20 | ···················· | ···················· | ····················· | ···················· | ···················· | ···················· |
| J | 3035 | hCoV-19/England/MILK-63BB9C/2020\|EPI_ISL_557784\|20 | ···················· | ···················· | ····················· | ···················· | ···················· | ···················· |
| J | 3036 | hCoV-19/England/ALDP-6B2A81/2020\|EPI_ISL_557785\|20 | ···················· | ···················· | ····················· | ···················· | ···················· | ···················· |
| J | 3037 | hCoV-19/England/ALDP-6C2C2E/2020\|EPI_ISL_557787\|20 | ···················· | ···················· | ····················· | ···················· | ···················· | ···················· |
| J | 3038 | hCoV-19/England/MILK-619095/2020\|EPI_ISL_557789\|20 | ···················· | ···················· | ····················· | ···················· | ···················· | ···················· |
| J | 3039 | hCoV-19/England/ALDP-6C3A41/2020\|EPI_ISL_557790\|20 | ···················· | ···················· | ····················· | ···················· | ···················· | ···················· |
| J | 3040 | hCoV-19/England/MILK-639509/2020\|EPI_ISL_557793\|20 | ···················· | ···················· | ····················· | ···················· | ···················· | ···················· |
| J | 3041 | hCoV-19/England/ALDP-6AFC90/2020\|EPI_ISL_557794\|20 | ···················· | ···················· | ····················· | ···················· | ···················· | ···················· |
| J | 3042 | hCoV-19/England/MILK-61CB28/2020\|EPI_ISL_557795\|20 | ···················· | ···················· | ····················· | ···················· | ···················· | ···················· |
| J | 3043 | hCoV-19/Wales/MILK-610878/2020\|EPI_ISL_557796\|2020 | ···················· | ···················· | ····················· | ···················· | ···················· | ···················· |
| J | 3044 | hCoV-19/England/ALDP-6B421E/2020\|EPI_ISL_557797\|20 | ···················· | ···················· | ····················· | ···················· | ···················· | ···················· |
| J | 3045 | hCoV-19/England/ALDP-6B580C/2020\|EPI_ISL_557798\|20 | ···················· | ···················· | ····················· | ···················· | ···················· | ···················· |
| J | 3046 | hCoV-19/England/ALDP-6BF2B9/2020\|EPI_ISL_557799\|20 | ···················· | ···················· | ····················· | ···················· | ···················· | ···················· |
| J | 3047 | hCoV-19/England/MILK-62AD4F/2020\|EPI_ISL_557800\|20 | ···················· | ···················· | ····················· | ···················· | ···················· | ···················· |
| J | 3048 | hCoV-19/England/MILK-625A10/2020\|EPI_ISL_557802\|20 | ···················· | ···················· | ····················· | ···················· | ···················· | ···················· |
| J | 3049 | hCoV-19/England/ALDP-6B8229/2020\|EPI_ISL_557803\|20 | ···················· | ···················· | ····················· | ···················· | ···················· | ···················· |
| J | 3050 | hCoV-19/England/ALDP-6B81E0/2020\|EPI_ISL_557804\|20 | ···················· | ···················· | ····················· | ···················· | ···················· | ···················· |
| J | 3051 | hCoV-19/England/ALDP-6C0CE4/2020\|EPI_ISL_557805\|20 | ···················· | ···················· | ····················· | ···················· | ···················· | ···················· |
| J | 3052 | hCoV-19/England/ALDP-6C1D95/2020\|EPI_ISL_557806\|20 | ···················· | ···················· | ····················· | ···················· | ···················· | ···················· |
| J | 3053 | hCoV-19/England/ALDP-6B4EED/2020\|EPI_ISL_557809\|20 | ···················· | ···················· | ····················· | ···················· | ···················· | ···················· |
| J | 3054 | hCoV-19/England/ALDP-6AEC46/2020\|EPI_ISL_557810\|20 | ···················· | ···················· | ····················· | ···················· | ···················· | ···················· |
| J | 3055 | hCoV-19/England/MILK-6166F0/2020\|EPI_ISL_557812\|20 | ···················· | ···················· | ···········T········· | ···················· | ···················· | ···················· |
| J | 3056 | hCoV-19/England/ALDP-6BBE5F/2020\|EPI_ISL_557813\|20 | ···················· | ···················· | ····················· | ···················· | ···················· | ···················· |
| J | 3057 | hCoV-19/England/ALDP-6B88BE/2020\|EPI_ISL_557814\|20 | ···················· | ···················· | ····················· | ···················· | ···················· | ···················· |
| J | 3058 | hCoV-19/England/ALDP-6B1D3E/2020\|EPI_ISL_557815\|20 | ···················· | ···················· | ····················· | ···················· | ···················· | ···················· |
| J | 3059 | hCoV-19/England/ALDP-6C091A/2020\|EPI_ISL_557818\|20 | ···················· | ···················· | ····················· | ···················· | ···················· | ···················· |
| J | 3060 | hCoV-19/England/ALDP-6B268D/2020\|EPI_ISL_557819\|20 | ···················· | ···················· | ····················· | ···················· | ···················· | ···················· |
| J | 3061 | hCoV-19/England/ALDP-6C1DE0/2020\|EPI_ISL_557820\|20 | ···················· | ···················· | ····················· | ···················· | ···················· | ···················· |
| J | 3062 | hCoV-19/England/MILK-628FB4/2020\|EPI_ISL_557821\|20 | ···················· | ···················· | ····················· | ···················· | ···················· | ···················· |
| J | 3063 | hCoV-19/England/MILK-62D447/2020\|EPI_ISL_557822\|20 | ···················· | ···················· | ····················· | ···················· | ···················· | ···················· |
| J | 3064 | hCoV-19/England/MILK-62D632/2020\|EPI_ISL_557823\|20 | ···················· | ···················· | ····················· | ···················· | ···················· | ···················· |
| J | 3065 | hCoV-19/England/MILK-60A237/2020\|EPI_ISL_557824\|20 | ···················· | ···················· | ····················· | ···················· | ···················· | ···················· |
| J | 3066 | hCoV-19/England/ALDP-6B59DC/2020\|EPI_ISL_557825\|20 | ···················· | ···················· | ····················· | ···················· | ···················· | ···················· |
| J | 3067 | hCoV-19/England/ALDP-6B53FC/2020\|EPI_ISL_557826\|20 | ···················· | ···················· | ····················· | ···················· | ···················· | ···················· |
| J | 3068 | hCoV-19/England/MILK-6145D7/2020\|EPI_ISL_557827\|20 | ···················· | ···················· | ····················· | ···················· | ···················· | ···················· |
| J | 3069 | hCoV-19/England/MILK-62BF1B/2020\|EPI_ISL_557829\|20 | ···················· | ···················· | ···········T········· | ···················· | ···················· | ···················· |
| J | 3070 | hCoV-19/England/MILK-615FF6/2020\|EPI_ISL_557830\|20 | ···················· | ···················· | ····················· | ···················· | ···················· | ···················· |
| J | 3071 | hCoV-19/England/ALDP-6BB693/2020\|EPI_ISL_557831\|20 | ···················· | ···················· | ····················· | ···················· | ···················· | ···················· |
| J | 3072 | hCoV-19/England/ALDP-6AD421/2020\|EPI_ISL_557833\|20 | ···················· | ···················· | ····················· | ···················· | ···················· | ···················· |
| J | 3073 | hCoV-19/England/MILK-9A8960/2020\|EPI_ISL_549329\|20 | ···················· | ···················· | ····················· | ···················· | ···················· | ···················· |
| J | 3074 | hCoV-19/England/MILK-9A8872/2020\|EPI_ISL_549333\|20 | ···················· | ···················· | ····················· | ···················· | ···················· | ···················· |
| J | 3075 | hCoV-19/England/NOTT-11336A/2020\|EPI_ISL_549334\|20 | ···················· | ···················· | ····················· | ···················· | ···················· | ···················· |
| J | 3076 | hCoV-19/England/NORW-EFB15/2020\|EPI_ISL_549336\|202 | ···················· | ···················· | ····················· | ···················· | ···················· | ···················· |
| J | 3077 | hCoV-19/England/MILK-9A8775/2020\|EPI_ISL_549342\|20 | ···················· | ···················· | ····················· | ···················· | ···················· | ···················· |
| J | 3078 | hCoV-19/England/MILK-9C4940/2020\|EPI_ISL_549343\|20 | ···················· | ···················· | ····················· | ···················· | ···················· | ···················· |
| J | 3079 | hCoV-19/England/NORW-EF68D/2020\|EPI_ISL_549344\|202 | ···················· | ···················· | ····················· | ···················· | ···················· | ···················· |
| J | 3080 | hCoV-19/England/QEUH-99632B/2020\|EPI_ISL_549345\|20 | ···················· | ···················· | ····················· | ···················· | ···················· | ···················· |
| J | 3081 | hCoV-19/England/QEUH-99672F/2020\|EPI_ISL_549346\|20 | ···················· | ···················· | ····················· | ···················· | ···················· | ···················· |
| J | 3082 | hCoV-19/England/CAMC-9B2338/2020\|EPI_ISL_549347\|20 | ···················· | ·A·················· | ····················· | ···················· | ···················· | ···A················ |
| J | 3083 | hCoV-19/England/MILK-9AB727/2020\|EPI_ISL_549348\|20 | ···················· | ·A·················· | ····················· | ···················· | ···················· | ···A················ |
| J | 3084 | hCoV-19/England/NORW-EFA45/2020\|EPI_ISL_549349\|202 | ···················· | ···················· | ····················· | ···················· | ···················· | ···················· |
| J | 3085 | hCoV-19/England/NORW-EF5DB/2020\|EPI_ISL_549350\|202 | ···················· | ···················· | ····················· | ···················· | ···················· | ···················· |
| J | 3086 | hCoV-19/England/NORW-EF67E/2020\|EPI_ISL_549351\|202 | ···················· | ···················· | ····················· | ···················· | ···················· | ···················· |
| J | 3087 | hCoV-19/England/MILK-99A7D0/2020\|EPI_ISL_550301\|20 | ···················· | ···················· | ····················· | ···················· | ···················· | ···················· |
| J | 3088 | hCoV-19/England/MILK-999FAC/2020\|EPI_ISL_550302\|20 | ···················· | ···················· | ····················· | ···················· | ···················· | ···················· |
| J | 3089 | hCoV-19/England/MILK-99A11E/2020\|EPI_ISL_550303\|20 | ···················· | ···················· | ····················· | ···················· | ···················· | ···················· |
| J | 3090 | hCoV-19/England/MILK-99A891/2020\|EPI_ISL_550304\|20 | ···················· | ·A·················· | ····················· | ···················· | ···················· | ···················· |
| J | 3091 | hCoV-19/England/MILK-99A3FA/2020\|EPI_ISL_550305\|20 | ···················· | ···················· | ····················· | ···················· | ···················· | ···················· |
| J | 3092 | hCoV-19/England/MILK-99A55E/2020\|EPI_ISL_550306\|20 | ···················· | ···················· | ····················· | ···················· | ···················· | ···················· |
| J | 3093 | hCoV-19/England/MILK-99B56C/2020\|EPI_ISL_550307\|20 | ···················· | ···················· | ····················· | ···················· | ···················· | ···················· |
| J | 3094 | hCoV-19/England/MILK-99A327/2020\|EPI_ISL_550308\|20 | ···················· | ·A·················· | ····················· | ···················· | ···················· | ···················· |
| J | 3095 | hCoV-19/England/MILK-999E45/2020\|EPI_ISL_550309\|20 | ···················· | ···················· | ····················· | ···················· | ···················· | ···················· |
| J | 3096 | hCoV-19/England/MILK-99A1D2/2020\|EPI_ISL_550310\|20 | ···················· | ···················· | ····················· | ···················· | ···················· | ···················· |
| J | 3097 | hCoV-19/England/MILK-999CB4/2020\|EPI_ISL_550311\|20 | ···················· | ···················· | ····················· | ···················· | ···················· | ···················· |

| | | | | | | | | |
|---|---|---|---|---|---|---|---|---|
| J | 3098 | hCoV-19/England/MILK-999FE8/2020\|EPI_ISL_550313\|20 | ···················· | ·A·················· | ···················· | ···················· | ···················· | ···················· |
| J | 3099 | hCoV-19/England/MILK-999BD5/2020\|EPI_ISL_550314\|20 | ···················· | ···················· | ···················· | ···················· | ···················· | ···················· |
| J | 3100 | hCoV-19/England/MILK-999F42/2020\|EPI_ISL_550315\|20 | ···················· | ···················· | ···················· | ···················· | ···················· | ···················· |
| J | 3101 | hCoV-19/England/MILK-999F8E/2020\|EPI_ISL_550316\|20 | ···················· | ···················· | ···················· | ···················· | ···················· | ···················· |
| J | 3102 | hCoV-19/England/MILK-99A882/2020\|EPI_ISL_550317\|20 | ···················· | ···················· | ···················· | ···················· | ···················· | ···················· |
| J | 3103 | hCoV-19/England/MILK-99A2B1/2020\|EPI_ISL_550318\|20 | ···················· | ···················· | ···················· | ···················· | ···················· | ···················· |
| J | 3104 | hCoV-19/Wales/MILK-99A08A/2020\|EPI_ISL_550319\|2020 | ···················· | ···················· | ···················· | ···················· | ···················· | ···················· |
| J | 3105 | hCoV-19/England/MILK-99A49D/2020\|EPI_ISL_550320\|20 | ···················· | ···················· | ···················· | ···················· | ···················· | ···················· |
| J | 3106 | hCoV-19/England/MILK-99A48E/2020\|EPI_ISL_550321\|20 | ···················· | ···················· | ···················· | ···················· | ···················· | ···················· |
| J | 3107 | hCoV-19/England/MILK-99A767/2020\|EPI_ISL_550323\|20 | ···················· | ···················· | ···················· | ···················· | ···················· | ···················· |
| J | 3108 | hCoV-19/England/MILK-99A12D/2020\|EPI_ISL_550324\|20 | ···················· | ···················· | ···················· | ···················· | ···················· | ···················· |
| J | 3109 | hCoV-19/England/MILK-99A169/2020\|EPI_ISL_550326\|20 | ···················· | ···················· | ···················· | ···················· | ···················· | ···················· |
| J | 3110 | hCoV-19/England/MILK-999C96/2020\|EPI_ISL_550327\|20 | ···················· | ···················· | ···················· | ···················· | ···················· | ···················· |
| J | 3111 | hCoV-19/Wales/MILK-99A6A6/2020\|EPI_ISL_550328\|2020 | ···················· | ···················· | ···················· | ···················· | ···················· | ···················· |
| J | 3112 | hCoV-19/England/MILK-99A9DA/2020\|EPI_ISL_550329\|20 | ···················· | ···················· | ···················· | ···················· | ···················· | ···················· |
| J | 3113 | hCoV-19/England/MILK-99A59A/2020\|EPI_ISL_550330\|20 | ···················· | ···················· | ···················· | ···················· | ···················· | ···················· |
| J | 3114 | hCoV-19/England/MILK-999F7F/2020\|EPI_ISL_550331\|20 | ···················· | ···················· | ···················· | ···················· | ···················· | ···················· |
| J | 3115 | hCoV-19/England/MILK-99A318/2020\|EPI_ISL_550337\|20 | ···················· | ···················· | ···················· | ···················· | ···················· | ···················· |
| J | 3116 | hCoV-19/England/MILK-99A0D5/2020\|EPI_ISL_550339\|20 | ···················· | ···················· | ···················· | ···················· | ···················· | ···················· |
| J | 3117 | hCoV-19/England/MILK-999E09/2020\|EPI_ISL_550340\|20 | ···················· | ···················· | ···················· | ···················· | ···················· | ···················· |
| J | 3118 | hCoV-19/England/MILK-99A187/2020\|EPI_ISL_550341\|20 | ···················· | ···················· | ···················· | ···················· | ···················· | ···················· |
| J | 3119 | hCoV-19/England/MILK-999F15/2020\|EPI_ISL_550342\|20 | ···················· | ···················· | ···················· | ···················· | ···················· | ···················· |
| J | 3120 | hCoV-19/England/MILK-999D57/2020\|EPI_ISL_550343\|20 | ···················· | ···················· | ···················· | ···················· | ···················· | ···················· |
| J | 3121 | hCoV-19/England/MILK-99A63D/2020\|EPI_ISL_550344\|20 | ···················· | ···················· | ···················· | ···················· | ···················· | ···················· |
| J | 3122 | hCoV-19/England/MILK-999F60/2020\|EPI_ISL_550345\|20 | ···················· | ···················· | ···················· | ···················· | ···················· | ···················· |
| J | 3123 | hCoV-19/England/MILK-99A8FB/2020\|EPI_ISL_550346\|20 | ···················· | ···················· | ···················· | ···················· | ···················· | ···················· |
| J | 3124 | hCoV-19/England/MILK-999C3C/2020\|EPI_ISL_550347\|20 | ···················· | ···················· | ···················· | ···················· | ···················· | ···················· |
| J | 3125 | hCoV-19/England/MILK-99A7FE/2020\|EPI_ISL_550348\|20 | ···················· | ···················· | ···················· | ···················· | ······T············· | ···················· |
| J | 3126 | hCoV-19/England/MILK-999E81/2020\|EPI_ISL_550349\|20 | ···················· | ···················· | ···················· | ···················· | ···················· | ···················· |
| J | 3127 | hCoV-19/England/MILK-99A0C6/2020\|EPI_ISL_550350\|20 | ···················· | ···················· | ···················· | ···················· | ···················· | ···················· |
| J | 3128 | hCoV-19/England/MILK-999EBE/2020\|EPI_ISL_550351\|20 | ···················· | ···················· | ···················· | ···················· | ···················· | ···················· |
| J | 3129 | hCoV-19/England/MILK-999EEB/2020\|EPI_ISL_550352\|20 | ···················· | ···················· | ···················· | ···················· | ···················· | ···················· |
| J | 3130 | hCoV-19/England/MILK-999FCA/2020\|EPI_ISL_550353\|20 | ···················· | ···················· | ···················· | ···················· | ···················· | ···················· |
| J | 3131 | hCoV-19/England/MILK-999C5A/2020\|EPI_ISL_550354\|20 | ···················· | ···················· | ···················· | ···················· | ···················· | ···················· |
| J | 3132 | hCoV-19/England/MILK-99A4D9/2020\|EPI_ISL_550355\|20 | ···················· | ···················· | ···················· | ···················· | ···················· | ···············A···· |
| J | 3133 | hCoV-19/England/MILK-99BB1F/2020\|EPI_ISL_550356\|20 | ···················· | ···················· | ···················· | ···················· | ···················· | ···················· |
| J | 3134 | hCoV-19/Wales/MILK-999CC3/2020\|EPI_ISL_550357\|2020 | ···················· | ···················· | ···················· | ···················· | ···················· | ···················· |
| J | 3135 | hCoV-19/England/MILK-99A1E1/2020\|EPI_ISL_550358\|20 | ···················· | ···················· | ···················· | ···················· | ···················· | ···················· |
| J | 3136 | hCoV-19/England/MILK-999C87/2020\|EPI_ISL_550359\|20 | ···················· | ···················· | ···················· | ···················· | ······T············· | ···················· |
| J | 3137 | hCoV-19/England/MILK-999D48/2020\|EPI_ISL_550360\|20 | ···················· | ···················· | ···················· | ···················· | ···················· | ···················· |
| J | 3138 | hCoV-19/England/MILK-999F06/2020\|EPI_ISL_550361\|20 | ···················· | ···················· | ···················· | ···················· | ···················· | ···················· |
| J | 3139 | hCoV-19/England/MILK-99A9BC/2020\|EPI_ISL_550362\|20 | ···················· | ···················· | ···················· | ···················· | ···················· | ···················· |
| J | 3140 | hCoV-19/England/MILK-99A47F/2020\|EPI_ISL_550363\|20 | ···················· | ···················· | ···················· | ···················· | ···················· | ···················· |
| J | 3141 | hCoV-19/Wales/MILK-99A06C/2020\|EPI_ISL_550364\|2020 | ···················· | ···················· | ···················· | ···················· | ···················· | ···················· |
| J | 3142 | hCoV-19/England/MILK-9943A5/2020\|EPI_ISL_551620\|20 | ···················· | ···················· | ···················· | ···················· | ···················· | ···················· |
| J | 3143 | hCoV-19/England/MILK-995984/2020\|EPI_ISL_551621\|20 | ···················· | ···················· | ···················· | ···················· | ···················· | ···················· |
| J | 3144 | hCoV-19/England/MILK-995AFA/2020\|EPI_ISL_551622\|20 | ···················· | ···················· | ···················· | ···················· | ···················· | ···················· |
| J | 3145 | hCoV-19/England/MILK-995975/2020\|EPI_ISL_551623\|20 | ···················· | ···················· | ···················· | ···················· | ···················· | ···················· |
| J | 3146 | hCoV-19/Wales/MILK-993F2A/2020\|EPI_ISL_551624\|2020 | ···················· | ···················· | ···················· | ···················· | ···················· | ···················· |
| J | 3147 | hCoV-19/England/MILK-99425D/2020\|EPI_ISL_551625\|20 | ···················· | ···················· | ···················· | ···················· | ···················· | ···················· |
| J | 3148 | hCoV-19/England/MILK-99568D/2020\|EPI_ISL_551626\|20 | ···················· | ···················· | ···················· | ···················· | ···················· | ···················· |
| J | 3149 | hCoV-19/England/MILK-9958B4/2020\|EPI_ISL_551629\|20 | ···················· | ···················· | ···················· | ···················· | ···················· | ···················· |
| J | 3150 | hCoV-19/England/MILK-9959FD/2020\|EPI_ISL_551632\|20 | ···················· | ···················· | ···················· | ···················· | ···················· | ···················· |
| J | 3151 | hCoV-19/England/MILK-994044/2020\|EPI_ISL_551633\|20 | ···················· | ·A·················· | ···················· | ···················· | ···················· | ···A················ |
| J | 3152 | hCoV-19/England/MILK-9942D5/2020\|EPI_ISL_551634\|20 | ···················· | ···················· | ···················· | ···················· | ···················· | ···················· |
| J | 3153 | hCoV-19/England/MILK-9958C3/2020\|EPI_ISL_551636\|20 | ···················· | ···················· | ···················· | ···················· | ···················· | ···················· |
| J | 3154 | hCoV-19/Wales/MILK-993DD5/2020\|EPI_ISL_551637\|2020 | ···················· | ···················· | ···················· | ···················· | ···················· | ···················· |
| J | 3155 | hCoV-19/England/MILK-99423F/2020\|EPI_ISL_551638\|20 | ···················· | ···················· | ···················· | ···················· | ···················· | ···················· |
| J | 3156 | hCoV-19/England/MILK-993E0F/2020\|EPI_ISL_551639\|20 | ···················· | ···················· | ···················· | ···················· | ···················· | ···················· |
| J | 3157 | hCoV-19/England/MILK-995887/2020\|EPI_ISL_551640\|20 | ···················· | ···················· | ···················· | ···················· | ···················· | ···················· |
| J | 3158 | hCoV-19/England/MILK-9957D5/2020\|EPI_ISL_551641\|20 | ···················· | ···················· | ···················· | ···················· | ···················· | ···················· |
| J | 3159 | hCoV-19/Wales/MILK-994220/2020\|EPI_ISL_551642\|2020 | ···················· | ···················· | ···················· | ···················· | ···················· | ···················· |
| J | 3160 | hCoV-19/England/MILK-995A36/2020\|EPI_ISL_551644\|20 | ···················· | ···················· | ···················· | ···················· | ···················· | ···················· |
| J | 3161 | hCoV-19/England/MILK-994062/2020\|EPI_ISL_551645\|20 | ···················· | ···················· | ···················· | ···················· | ···················· | ···················· |
| J | 3162 | hCoV-19/England/MILK-9941D8/2020\|EPI_ISL_551646\|20 | ···················· | ······A············· | ···················· | ···················· | ···················· | ···················· |
| J | 3163 | hCoV-19/England/MILK-99578A/2020\|EPI_ISL_551648\|20 | ···················· | ·A·················· | ···················· | ···················· | ···················· | ···················· |
| J | 3164 | hCoV-19/Wales/MILK-993F75/2020\|EPI_ISL_551649\|2020 | ···················· | ···················· | ···················· | ···················· | ···················· | ···················· |
| J | 3165 | hCoV-19/England/MILK-995ACD/2020\|EPI_ISL_551650\|20 | ···················· | ···················· | ···················· | ···················· | ···················· | ···················· |
| J | 3166 | hCoV-19/England/MILK-993EF0/2020\|EPI_ISL_551651\|20 | ········T··········· | ···················· | ···················· | ···················· | ···················· | ···················· |
| J | 3167 | hCoV-19/England/MILK-994387/2020\|EPI_ISL_551653\|20 | ···················· | ···················· | ···················· | ···················· | ···················· | ···················· |
| J | 3168 | hCoV-19/England/MILK-9940DB/2020\|EPI_ISL_551654\|20 | ···················· | ···················· | ···················· | ···················· | ···················· | ···················· |
| J | 3169 | hCoV-19/England/MILK-994448/2020\|EPI_ISL_551655\|20 | ···················· | ···················· | ···················· | ···················· | ···················· | ···················· |
| J | 3170 | hCoV-19/England/MILK-9940EA/2020\|EPI_ISL_551656\|20 | ···················· | ···················· | ···················· | ···················· | ···················· | ···················· |
| J | 3171 | hCoV-19/England/MILK-994475/2020\|EPI_ISL_551657\|20 | ···················· | ···················· | ···················· | ···················· | ···················· | ···················· |
| J | 3172 | hCoV-19/England/MILK-9957B7/2020\|EPI_ISL_551658\|20 | ···················· | ···················· | ···················· | ···················· | ···················· | ···················· |
| J | 3173 | hCoV-19/England/MILK-995C9A/2020\|EPI_ISL_551661\|20 | ···················· | ···················· | ···················· | ···················· | ···················· | ···················· |
| J | 3174 | hCoV-19/England/MILK-994053/2020\|EPI_ISL_551662\|20 | ···················· | ···················· | ···················· | ···················· | ···················· | ···················· |
| J | 3175 | hCoV-19/England/MILK-99417E/2020\|EPI_ISL_551664\|20 | ···················· | ···················· | ···················· | ···················· | ···················· | ···················· |
| J | 3176 | hCoV-19/England/MILK-99432D/2020\|EPI_ISL_551665\|20 | ···················· | ···················· | ···················· | ···················· | ···················· | ···················· |
| J | 3177 | hCoV-19/England/MILK-9941C9/2020\|EPI_ISL_551667\|20 | ···················· | ···················· | ···················· | ···················· | ···················· | ···················· |

| | | | | | | | | |
|---|---|---|---|---|---|---|---|---|
| J | 3178 | hCoV-19/England/MILK-993FC0/2020\|EPI_ISL_551670\|20 | ···················· | ···················· | ····················· | ····················· | ···················· | ···················· |
| J | 3179 | hCoV-19/England/MILK-995939/2020\|EPI_ISL_551671\|20 | ···················· | ···················· | ····················· | ····················· | ···················· | ···················· |
| J | 3180 | hCoV-19/England/MILK-993E4B/2020\|EPI_ISL_551672\|20 | ···················· | ···················· | ····················· | ····················· | ···················· | ···················· |
| J | 3181 | hCoV-19/England/MILK-9942F3/2020\|EPI_ISL_551674\|20 | ···················· | ···················· | ····················· | ····················· | ···················· | ···················· |
| J | 3182 | hCoV-19/England/MILK-994114/2020\|EPI_ISL_551675\|20 | ···················· | ·A·················· | ····················· | ····················· | ···················· | ··A················· |
| J | 3183 | hCoV-19/England/MILK-993F93/2020\|EPI_ISL_551676\|20 | ···················· | ···················· | ····················· | ····················· | ···················· | ···················· |
| J | 3184 | hCoV-19/England/MILK-993ED2/2020\|EPI_ISL_551677\|20 | ···················· | ···················· | ····················· | ····················· | ···················· | ···················· |
| J | 3185 | hCoV-19/England/MILK-9942C6/2020\|EPI_ISL_551678\|20 | ···················· | ···················· | ····················· | ····················· | ···················· | ···················· |
| J | 3186 | hCoV-19/England/MILK-995993/2020\|EPI_ISL_551680\|20 | ···················· | ···················· | ····················· | ····················· | ···················· | ···················· |
| J | 3187 | hCoV-19/England/MILK-995544/2020\|EPI_ISL_551683\|20 | ···················· | ···················· | ····················· | ····················· | ···················· | ···················· |
| J | 3188 | hCoV-19/England/MILK-993E5A/2020\|EPI_ISL_551684\|20 | ···················· | ···················· | ····················· | ····················· | ···················· | ···················· |
| J | 3189 | hCoV-19/England/MILK-993E96/2020\|EPI_ISL_551685\|20 | ···················· | ···················· | ····················· | ····················· | ···················· | ···················· |
| J | 3190 | hCoV-19/England/MILK-99577B/2020\|EPI_ISL_551687\|20 | ···················· | ···················· | ····················· | ····················· | ···················· | ···················· |
| J | 3191 | hCoV-19/England/MILK-9958F0/2020\|EPI_ISL_551688\|20 | ···················· | ···················· | ····················· | ····················· | ···················· | ···················· |
| J | 3192 | hCoV-19/England/MILK-9956AB/2020\|EPI_ISL_551689\|20 | ···················· | ···················· | ····················· | ····················· | ···················· | ···················· |
| J | 3193 | hCoV-19/England/MILK-995948/2020\|EPI_ISL_551690\|20 | ···················· | ···················· | ····················· | ····················· | ···················· | ···················· |
| J | 3194 | hCoV-19/England/MILK-993FEE/2020\|EPI_ISL_551691\|20 | ···················· | ···················· | ····················· | ····················· | ···················· | ···················· |
| J | 3195 | hCoV-19/Wales/MILK-993F66/2020\|EPI_ISL_551692\|2020 | ···················· | ···················· | ····················· | ····················· | ···················· | ···················· |
| J | 3196 | hCoV-19/England/MILK-995702/2020\|EPI_ISL_551695\|20 | ···················· | ···················· | ····················· | ····················· | ···················· | ···················· |
| J | 3197 | hCoV-19/England/MILK-93C0B7/2020\|EPI_ISL_552950\|20 | ···················· | ···················· | ····················· | ····················· | ···················· | ···················· |
| J | 3198 | hCoV-19/England/ALDP-931FC4/2020\|EPI_ISL_552951\|20 | ···················· | ···················· | ····················· | ····················· | ···················· | ···················· |
| J | 3199 | hCoV-19/Wales/MILK-93AF16/2020\|EPI_ISL_552953\|2020 | ···················· | ···················· | ····················· | ····················· | ···················· | ···················· |
| J | 3200 | hCoV-19/England/ALDP-92A966/2020\|EPI_ISL_552954\|20 | ···················· | ···················· | ····················· | ····················· | ···················· | ···················· |
| J | 3201 | hCoV-19/England/ALDP-92933C/2020\|EPI_ISL_552955\|20 | ···················· | ···················· | ····················· | ····················· | ···················· | ···················· |
| J | 3202 | hCoV-19/England/ALDP-935555/2020\|EPI_ISL_552956\|20 | ···················· | ···················· | ····················· | ····················· | ···················· | ···················· |
| J | 3203 | hCoV-19/England/ALDP-939ECF/2020\|EPI_ISL_552957\|20 | ···················· | ···················· | ····················· | ····················· | ···················· | ···················· |
| J | 3204 | hCoV-19/England/ALDP-932084/2020\|EPI_ISL_552958\|20 | ···················· | ···················· | ····················· | ····················· | ···················· | ···················· |
| J | 3205 | hCoV-19/England/ALDP-933BBF/2020\|EPI_ISL_552964\|20 | ···················· | ···················· | ····················· | ····················· | ···················· | ···················· |
| J | 3206 | hCoV-19/England/ALDP-929369/2020\|EPI_ISL_552965\|20 | ···················· | ···················· | ····················· | ····················· | ···················· | ···················· |
| J | 3207 | hCoV-19/England/ALDP-92B3C1/2020\|EPI_ISL_552966\|20 | ···················· | ···················· | ····················· | ····················· | ···················· | ···················· |
| J | 3208 | hCoV-19/England/ALDP-92B5F8/2020\|EPI_ISL_552968\|20 | ···················· | ···················· | ····················· | ····················· | ···················· | ···················· |
| J | 3209 | hCoV-19/England/ALDP-92CAE9/2020\|EPI_ISL_552969\|20 | ···················· | ···················· | ····················· | ····················· | ···················· | ···················· |
| J | 3210 | hCoV-19/Wales/ALDP-9362B7/2020\|EPI_ISL_552970\|2020 | ···················· | ···················· | ····················· | ····················· | ···················· | ···················· |
| J | 3211 | hCoV-19/England/ALDP-920450/2020\|EPI_ISL_552971\|20 | ···················· | ···················· | ····················· | ····················· | ···················· | ···················· |
| J | 3212 | hCoV-19/Wales/ALDP-920C2B/2020\|EPI_ISL_552974\|2020 | ···················· | ·A·················· | ····················· | ····················· | ···················· | ···················· |
| J | 3213 | hCoV-19/England/ALDP-913CFE/2020\|EPI_ISL_552976\|20 | ···················· | ···················· | ····················· | ····················· | ···················· | ···················· |
| J | 3214 | hCoV-19/England/ALDP-935D8A/2020\|EPI_ISL_552977\|20 | ···················· | ·A·················· | ····················· | ····················· | ···················· | ···················· |
| J | 3215 | hCoV-19/England/ALDP-934802/2020\|EPI_ISL_552978\|20 | ···················· | ···················· | ····················· | ····················· | ···················· | ···················· |
| J | 3216 | hCoV-19/England/MILK-93C828/2020\|EPI_ISL_552979\|20 | ···················· | ···················· | ····················· | ····················· | ···················· | ···················· |
| J | 3217 | hCoV-19/England/ALDP-928B71/2020\|EPI_ISL_552980\|20 | ···················· | ···················· | ····················· | ····················· | ···················· | ···················· |
| J | 3218 | hCoV-19/England/ALDP-9241CE/2020\|EPI_ISL_552981\|20 | ···················· | ···················· | ····················· | ····················· | ···················· | ···················· |
| J | 3219 | hCoV-19/Wales/ALDP-919C9E/2020\|EPI_ISL_552982\|2020 | ···················· | ···················· | ····················· | ····················· | ···················· | ···················· |
| J | 3220 | hCoV-19/England/ALDP-9038DF/2020\|EPI_ISL_552983\|20 | ···················· | ···················· | ····················· | ····················· | ···················· | ···················· |
| J | 3221 | hCoV-19/England/ALDP-93153B/2020\|EPI_ISL_552984\|20 | ···················· | ·A·················· | ····················· | ····················· | ···················· | ···················· |
| J | 3222 | hCoV-19/England/ALDP-929080/2020\|EPI_ISL_552985\|20 | ···················· | ···················· | ····················· | ····················· | ···················· | ···················· |
| J | 3223 | hCoV-19/England/ALDP-920A6D/2020\|EPI_ISL_552986\|20 | ···················· | ·A·················· | ····················· | ····················· | ···················· | ···················· |
| J | 3224 | hCoV-19/England/MILK-938561/2020\|EPI_ISL_552987\|20 | ···················· | ···················· | ····················· | ····················· | ···················· | ···················· |
| J | 3225 | hCoV-19/England/ALDP-933A76/2020\|EPI_ISL_552988\|20 | ···················· | ···················· | ····················· | ····················· | ···················· | ···················· |
| J | 3226 | hCoV-19/England/ALDP-92887A/2020\|EPI_ISL_552989\|20 | ···················· | ···················· | ····················· | ····················· | ···················· | ···················· |
| J | 3227 | hCoV-19/England/MILK-93F2BE/2020\|EPI_ISL_552990\|20 | ···················· | ···················· | ····················· | ····················· | ···················· | ···················· |
| J | 3228 | hCoV-19/England/ALDP-929448/2020\|EPI_ISL_552991\|20 | ···················· | ···················· | ····················· | ····················· | ···················· | ···················· |
| J | 3229 | hCoV-19/England/ALDP-926D22/2020\|EPI_ISL_552992\|20 | ···················· | ···················· | ····················· | ····················· | ···················· | ···················· |
| J | 3230 | hCoV-19/England/ALDP-928FEE/2020\|EPI_ISL_552993\|20 | ···················· | ···················· | ····················· | ····················· | ···················· | ···················· |
| J | 3231 | hCoV-19/England/ALDP-933830/2020\|EPI_ISL_552994\|20 | ···················· | ···················· | ····················· | ····················· | ···················· | ···················· |
| J | 3232 | hCoV-19/England/MILK-93A1E3/2020\|EPI_ISL_552995\|20 | ···················· | ···················· | ····················· | ····················· | ···················· | ···················· |
| J | 3233 | hCoV-19/England/ALDP-90320E/2020\|EPI_ISL_552996\|20 | ···················· | ···················· | ····················· | ····················· | ···················· | ···················· |
| J | 3234 | hCoV-19/England/MILK-938640/2020\|EPI_ISL_552997\|20 | ···················· | ···················· | ····················· | ····················· | ···················· | ···················· |
| J | 3235 | hCoV-19/England/ALDP-92693D/2020\|EPI_ISL_552998\|20 | ···················· | ···················· | ····················· | ····················· | ···················· | ···················· |
| J | 3236 | hCoV-19/England/ALDP-931735/2020\|EPI_ISL_552999\|20 | ···················· | ···················· | ····················· | ····················· | ···················· | ···················· |
| J | 3237 | hCoV-19/England/ALDP-94D88E/2020\|EPI_ISL_555021\|20 | ···················· | ···················· | ····················· | ····················· | ···················· | ···················· |
| J | 3238 | hCoV-19/England/ALDP-95C50D/2020\|EPI_ISL_555022\|20 | ···················· | ···················· | ····················· | ····················· | ···················· | ···················· |
| J | 3239 | hCoV-19/England/ALDP-94E95D/2020\|EPI_ISL_555024\|20 | ···················· | ···················· | ····················· | ····················· | ···················· | ···················· |
| J | 3240 | hCoV-19/England/ALDP-94D754/2020\|EPI_ISL_555025\|20 | ···················· | ···················· | ····················· | ····················· | ···················· | ···················· |
| J | 3241 | hCoV-19/England/ALDP-94D736/2020\|EPI_ISL_555026\|20 | ···················· | ·A·················· | ····················· | ····················· | ···················· | ···················· |
| J | 3242 | hCoV-19/England/ALDP-95C075/2020\|EPI_ISL_555027\|20 | ···················· | ···················· | ····················· | ····················· | ···················· | ···················· |
| J | 3243 | hCoV-19/England/ALDP-95A62A/2020\|EPI_ISL_555028\|20 | ···················· | ·A·················· | ····················· | ····················· | ···················· | ···················· |
| J | 3244 | hCoV-19/England/ALDP-959EE7/2020\|EPI_ISL_555029\|20 | ···················· | ···················· | ···········T········· | ····················· | ···················· | ···················· |
| J | 3245 | hCoV-19/England/ALDP-94D4D5/2020\|EPI_ISL_555030\|20 | ···················· | ···················· | ····················· | ····················· | ···················· | ···················· |
| J | 3246 | hCoV-19/England/ALDP-95C840/2020\|EPI_ISL_555032\|20 | ···················· | ···················· | ····················· | ····················· | ···················· | ···················· |
| J | 3247 | hCoV-19/England/ALDP-95C206/2020\|EPI_ISL_555033\|20 | ···················· | ···················· | ····················· | ····················· | ···················· | ···················· |
| J | 3248 | hCoV-19/England/ALDP-95C51C/2020\|EPI_ISL_555034\|20 | ···················· | ···················· | ····················· | ····················· | ···················· | ···················· |
| J | 3249 | hCoV-19/England/ALDP-95C6EC/2020\|EPI_ISL_555035\|20 | ···················· | ···················· | ····················· | ····················· | ···················· | ···················· |
| J | 3250 | hCoV-19/England/ALDP-95C34F/2020\|EPI_ISL_555037\|20 | ···················· | ···················· | ····················· | ····················· | ···················· | ···················· |
| J | 3251 | hCoV-19/England/ALDP-95C37C/2020\|EPI_ISL_555038\|20 | ···················· | ···················· | ····················· | ····················· | ···················· | ···················· |
| J | 3252 | hCoV-19/England/ALDP-94E94E/2020\|EPI_ISL_555039\|20 | ···················· | ···················· | ····················· | ····················· | ···················· | ···················· |
| J | 3253 | hCoV-19/England/ALDP-95A226/2020\|EPI_ISL_555040\|20 | ···················· | ···················· | ····················· | ····················· | ···················· | ···················· |
| J | 3254 | hCoV-19/England/ALDP-95A7BE/2020\|EPI_ISL_555042\|20 | ···················· | ···················· | ····················· | ····················· | ···················· | ···················· |
| J | 3255 | hCoV-19/England/ALDP-959D35/2020\|EPI_ISL_555043\|20 | ···················· | ···················· | ····················· | ····················· | ···················· | ···················· |
| J | 3256 | hCoV-19/England/ALDP-94D5D2/2020\|EPI_ISL_555044\|20 | ···················· | ···················· | ····················· | ····················· | ···················· | ···················· |
| J | 3257 | hCoV-19/England/ALDP-94D62A/2020\|EPI_ISL_555045\|20 | ···················· | ···················· | ····················· | ····················· | ···················· | ···················· |

| | | | | | | | |
|---|---|---|---|---|---|---|---|
| J | 3258 hCoV-19/England/ALDP-95C585/2020\|EPI_ISL_555046\|20 | ···················· | ···················· | ····················· | ···················· | ···················· | ···················· |
| J | 3259 hCoV-19/England/ALDP-94D675/2020\|EPI_ISL_555047\|20 | ···················· | ···················· | ····················· | ···················· | ···················· | ···················· |
| J | 3260 hCoV-19/England/ALDP-94E841/2020\|EPI_ISL_555048\|20 | ···················· | ···················· | ····················· | ···················· | ···················· | ···················· |
| J | 3261 hCoV-19/England/ALDP-94EA78/2020\|EPI_ISL_555049\|20 | ···················· | ···················· | ····················· | ···················· | ···················· | ···················· |
| J | 3262 hCoV-19/England/ALDP-94D6EE/2020\|EPI_ISL_555050\|20 | ···················· | ···················· | ····················· | ···················· | ···················· | ···················· |
| J | 3263 hCoV-19/England/ALDP-95C53A/2020\|EPI_ISL_555051\|20 | ···················· | ···················· | ····················· | ···················· | ···················· | ···················· |
| J | 3264 hCoV-19/England/ALDP-95A9C7/2020\|EPI_ISL_555052\|20 | ···················· | ·A·················· | ····················· | ···················· | ···················· | ···················· |
| J | 3265 hCoV-19/England/ALDP-94E79F/2020\|EPI_ISL_555053\|20 | ···················· | ···················· | ····················· | ···················· | ···················· | ···················· |
| J | 3266 hCoV-19/England/ALDP-95C190/2020\|EPI_ISL_555054\|20 | ···················· | ···················· | ····················· | ···················· | ···················· | ···················· |
| J | 3267 hCoV-19/England/ALDP-95C35E/2020\|EPI_ISL_555055\|20 | ···················· | ···················· | ····················· | ···················· | ···················· | ···················· |
| J | 3268 hCoV-19/England/ALDP-94D772/2020\|EPI_ISL_555057\|20 | ···················· | ···················· | ····················· | ···················· | ···················· | ···················· |
| J | 3269 hCoV-19/England/ALDP-95C3C7/2020\|EPI_ISL_555058\|20 | ···················· | ···················· | ····················· | ···················· | ···················· | ···················· |
| J | 3270 hCoV-19/England/ALDP-94D6B1/2020\|EPI_ISL_555059\|20 | ···················· | ···················· | ····················· | ···················· | ···················· | ···················· |
| J | 3271 hCoV-19/England/ALDP-94E780/2020\|EPI_ISL_555060\|20 | ···················· | ···················· | ····················· | ···················· | ···················· | ···················· |
| J | 3272 hCoV-19/England/ALDP-94D4E4/2020\|EPI_ISL_555061\|20 | ···················· | ···················· | ····················· | ···················· | ···················· | ···················· |
| J | 3273 hCoV-19/Wales/ALDP-95AAE2/2020\|EPI_ISL_555064\|2020 | ···················· | ···················· | ····················· | ···················· | ···················· | ···················· |
| J | 3274 hCoV-19/England/ALDP-95C8AA/2020\|EPI_ISL_555065\|20 | ···················· | ···················· | ····················· | ···················· | ···················· | ···················· |
| J | 3275 hCoV-19/England/ALDP-94E93F/2020\|EPI_ISL_555066\|20 | ···················· | ···················· | ····················· | ···················· | ···················· | ···················· |
| J | 3276 hCoV-19/England/ALDP-95C497/2020\|EPI_ISL_555067\|20 | ···················· | ···················· | ····················· | ···················· | ···················· | ···················· |
| J | 3277 hCoV-19/England/ALDP-94E814/2020\|EPI_ISL_555069\|20 | ···················· | ···················· | ····················· | ···················· | ···················· | ···················· |
| J | 3278 hCoV-19/England/ALDP-94E920/2020\|EPI_ISL_555078\|20 | ···················· | ···················· | ····················· | ···················· | ···················· | ···················· |
| J | 3279 hCoV-19/England/CAMC-949F9C/2020\|EPI_ISL_556351\|20 | ···················· | ···················· | ···········T········· | ···················· | ···················· | ···················· |
| J | 3280 hCoV-19/England/ALDP-9498A0/2020\|EPI_ISL_556352\|20 | ···················· | ···················· | ····················· | ···················· | ···················· | ···················· |
| J | 3281 hCoV-19/England/ALDP-94A4F6/2020\|EPI_ISL_556355\|20 | ···················· | ···················· | ····················· | ···················· | ···················· | ···················· |
| J | 3282 hCoV-19/England/ALDP-94A71B/2020\|EPI_ISL_556356\|20 | ···················· | ···················· | ····················· | ···················· | ···················· | ···················· |
| J | 3283 hCoV-19/England/CAMC-94B64A/2020\|EPI_ISL_556357\|20 | ···················· | ···················· | ····················· | ···················· | ···················· | ···················· |
| J | 3284 hCoV-19/England/ALDP-94A07A/2020\|EPI_ISL_556358\|20 | ···················· | ···················· | ····················· | ···················· | ···················· | ···················· |
| J | 3285 hCoV-19/England/ALDP-949D65/2020\|EPI_ISL_556359\|20 | ···················· | ···················· | ····················· | ···················· | ···················· | ···················· |
| J | 3286 hCoV-19/England/ALDP-94EF97/2020\|EPI_ISL_556360\|20 | ···················· | ···················· | ····················· | ···················· | ···················· | ···················· |
| J | 3287 hCoV-19/England/ALDP-94F181/2020\|EPI_ISL_556361\|20 | ···················· | ···················· | ····················· | ···················· | ···················· | ···················· |
| J | 3288 hCoV-19/England/ALDP-94A177/2020\|EPI_ISL_556362\|20 | ···················· | ···················· | ····················· | ···················· | ···················· | ···················· |
| J | 3289 hCoV-19/England/ALDP-94A06B/2020\|EPI_ISL_556365\|20 | ···················· | ···················· | ····················· | ···················· | ···················· | ···················· |
| J | 3290 hCoV-19/England/ALDP-94EBA2/2020\|EPI_ISL_556366\|20 | ···················· | ·A·················· | ····················· | ···················· | ···················· | ···················· |
| J | 3291 hCoV-19/England/ALDP-94D244/2020\|EPI_ISL_556367\|20 | ···················· | ·A·················· | ····················· | ···················· | ···················· | ···················· |
| J | 3292 hCoV-19/England/ALDP-94A57B/2020\|EPI_ISL_556369\|20 | ···················· | ···················· | ····················· | ···················· | ···················· | ···················· |
| J | 3293 hCoV-19/England/ALDP-94A362/2020\|EPI_ISL_556370\|20 | ···················· | ···················· | ····················· | ···················· | ···················· | ···················· |
| J | 3294 hCoV-19/England/ALDP-94F163/2020\|EPI_ISL_556373\|20 | ···················· | ···················· | ····················· | ···················· | ···················· | ···················· |
| J | 3295 hCoV-19/England/ALDP-94A4E7/2020\|EPI_ISL_556375\|20 | ···················· | ···················· | ····················· | ···················· | ···················· | ···················· |
| J | 3296 hCoV-19/England/ALDP-94EC63/2020\|EPI_ISL_556376\|20 | ···················· | ···················· | ····················· | ···················· | ···················· | ···················· |
| J | 3297 hCoV-19/England/ALDP-94F707/2020\|EPI_ISL_556377\|20 | ···················· | ···················· | ····················· | ···················· | ···················· | ···················· |
| J | 3298 hCoV-19/England/ALDP-94AF22/2020\|EPI_ISL_556379\|20 | ···················· | ···················· | ····················· | ···················· | ···················· | ···················· |
| J | 3299 hCoV-19/England/ALDP-94ECFA/2020\|EPI_ISL_556380\|20 | ···················· | ···················· | ····················· | ···················· | ···················· | ···················· |
| J | 3300 hCoV-19/England/ALDP-94A195/2020\|EPI_ISL_556381\|20 | ···················· | ·A·················· | ····················· | ···················· | ···················· | ···················· |
| J | 3301 hCoV-19/England/ALDP-94A58A/2020\|EPI_ISL_556384\|20 | ···················· | ···················· | ····················· | ···················· | ···················· | ···················· |
| J | 3302 hCoV-19/England/ALDP-94ECEB/2020\|EPI_ISL_556385\|20 | ···················· | ···················· | ····················· | ···················· | ···················· | ···················· |
| J | 3303 hCoV-19/England/ALDP-94D174/2020\|EPI_ISL_556386\|20 | ···················· | ···················· | ····················· | ···················· | ···················· | ···················· |
| J | 3304 hCoV-19/England/ALDP-94EDE8/2020\|EPI_ISL_556387\|20 | ···················· | ···················· | ····················· | ···················· | ···················· | ···················· |
| J | 3305 hCoV-19/England/ALDP-94AEDA/2020\|EPI_ISL_556389\|20 | ···················· | ···················· | ····················· | ···················· | ···················· | ···················· |
| J | 3306 hCoV-19/England/ALDP-94AF40/2020\|EPI_ISL_556390\|20 | ···················· | ···················· | ····················· | ···················· | ···················· | ···················· |
| J | 3307 hCoV-19/England/ALDP-94B8EA/2020\|EPI_ISL_556391\|20 | ···················· | ···················· | ····················· | ···················· | ···················· | ···················· |
| J | 3308 hCoV-19/England/ALDP-94EDAC/2020\|EPI_ISL_556392\|20 | ···················· | ···················· | ····················· | ···················· | ···················· | ···················· |
| J | 3309 hCoV-19/Wales/ALDP-94EE30/2020\|EPI_ISL_556393\|2020 | ···················· | ···················· | ····················· | ···················· | ···················· | ···················· |
| J | 3310 hCoV-19/England/ALDP-9498CE/2020\|EPI_ISL_556394\|20 | ···················· | ···················· | ····················· | ···················· | ···················· | ···················· |
| J | 3311 hCoV-19/England/ALDP-94A04D/2020\|EPI_ISL_556395\|20 | ···················· | ···················· | ····················· | ···················· | ···················· | ···················· |
| J | 3312 hCoV-19/England/ALDP-94F0A2/2020\|EPI_ISL_556396\|20 | ···················· | ···················· | ····················· | ···················· | ···················· | ···················· |
| J | 3313 hCoV-19/England/ALDP-94D350/2020\|EPI_ISL_556397\|20 | ···················· | ···················· | ····················· | ···················· | ···················· | ···················· |
| J | 3314 hCoV-19/England/ALDP-94AF5F/2020\|EPI_ISL_556398\|20 | ···················· | ···················· | ····················· | ···················· | ···················· | ···················· |
| J | 3315 hCoV-19/England/ALDP-94EFA6/2020\|EPI_ISL_556399\|20 | ···················· | ···················· | ····················· | ···················· | ···················· | ···················· |
| J | 3316 hCoV-19/England/ALDP-7091C1/2020\|EPI_ISL_557682\|20 | ···················· | ···················· | ····················· | ···················· | ···················· | ···················· |
| J | 3317 hCoV-19/England/ALDP-722FB9/2020\|EPI_ISL_557683\|20 | ···················· | ···················· | ····················· | ···················· | ···················· | ···················· |
| J | 3318 hCoV-19/England/ALDP-722A6D/2020\|EPI_ISL_557684\|20 | ···················· | ···················· | ····················· | ···················· | ···················· | ···················· |
| J | 3319 hCoV-19/England/ALDP-724C0B/2020\|EPI_ISL_557686\|20 | ···················· | ···················· | ····················· | ···················· | ···················· | ···················· |
| J | 3320 hCoV-19/England/ALDP-725B67/2020\|EPI_ISL_557687\|20 | ···················· | ···················· | ····················· | ···················· | ···················· | ···················· |
| J | 3321 hCoV-19/England/ALDP-723598/2020\|EPI_ISL_557688\|20 | ···················· | ···················· | ····················· | ···················· | ···················· | ···················· |
| J | 3322 hCoV-19/England/ALDP-71C547/2020\|EPI_ISL_557690\|20 | ···················· | ···················· | ····················· | ···················· | ···················· | ···················· |
| J | 3323 hCoV-19/England/ALDP-71DF75/2020\|EPI_ISL_557691\|20 | ···················· | ···················· | ····················· | ···················· | ···················· | ···················· |
| J | 3324 hCoV-19/England/ALDP-6E825D/2020\|EPI_ISL_557693\|20 | ···················· | ···················· | ····················· | ···················· | ···················· | ···················· |
| J | 3325 hCoV-19/England/ALDP-721415/2020\|EPI_ISL_557695\|20 | ···················· | ···················· | ····················· | ···················· | ···················· | ···················· |
| J | 3326 hCoV-19/England/ALDP-7090A6/2020\|EPI_ISL_557697\|20 | ···················· | ·A·················· | ····················· | ···················· | ···················· | ···················· |
| J | 3327 hCoV-19/England/ALDP-725727/2020\|EPI_ISL_557698\|20 | ···················· | ···················· | ····················· | ···················· | ···················· | ···················· |
| J | 3328 hCoV-19/England/ALDP-704B5F/2020\|EPI_ISL_557699\|20 | ···················· | ···················· | ····················· | ···················· | ···················· | ···················· |
| J | 3329 hCoV-19/USA/FL-BPHL-1480/2020\|EPI_ISL_549262\|2020- | ···················· | ···················· | ····················· | ···················· | ···················· | ···················· |
| J | 3330 hCoV-19/USA/FL-BPHL-1482/2020\|EPI_ISL_549264\|2020- | ···················· | ···················· | ····················· | ···················· | ···················· | ···················· |
| J | 3331 hCoV-19/USA/FL-BPHL-1483/2020\|EPI_ISL_549265\|2020- | ···················· | ···················· | ····················· | ···················· | ···················· | ···················· |
| J | 3332 hCoV-19/USA/FL-BPHL-1484/2020\|EPI_ISL_549266\|2020- | ···················· | ···················· | ····················· | ···················· | ···················· | ···················· |
| J | 3333 hCoV-19/USA/FL-BPHL-1485/2020\|EPI_ISL_549267\|2020- | ···················· | ···················· | ····················· | ···················· | ···················· | ···················· |
| J | 3334 hCoV-19/USA/FL-BPHL-1486/2020\|EPI_ISL_549268\|2020- | ···················· | ···················· | ····················· | ···················· | ···················· | ···················· |
| J | 3335 hCoV-19/USA/FL-BPHL-1487/2020\|EPI_ISL_549269\|2020- | ···················· | ···················· | ····················· | ···················· | ···················· | ···················· |
| J | 3336 hCoV-19/Wales/MILK-9A53ED/2020\|EPI_ISL_550244\|2020 | ···················· | ···················· | ····················· | ···················· | ···················· | ···················· |
| J | 3337 hCoV-19/England/MILK-9A6F24/2020\|EPI_ISL_550245\|20 | ···················· | ···················· | ····················· | ···················· | ···················· | ···················· |

| | | | | | | | | |
|---|---|---|---|---|---|---|---|---|
| J | 3338 | hCoV-19/England/MILK-9A69BD/2020\|EPI_ISL_550246\|20 | ···················· | ···················· | ····················· | ···················· | ···················· | ···················· |
| J | 3339 | hCoV-19/England/MILK-9A82FC/2020\|EPI_ISL_550247\|20 | ···················· | ···················· | ····················· | ···················· | ···················· | ···················· |
| J | 3340 | hCoV-19/England/MILK-9A6917/2020\|EPI_ISL_550248\|20 | ···················· | ···················· | ····················· | ···················· | ···················· | ···················· |
| J | 3341 | hCoV-19/England/MILK-9A6D48/2020\|EPI_ISL_550249\|20 | ···················· | ···················· | ····················· | ···················· | ···················· | ···················· |
| J | 3342 | hCoV-19/England/MILK-9A860F/2020\|EPI_ISL_550250\|20 | ···················· | ···················· | ····················· | ···················· | ···················· | ···················· |
| J | 3343 | hCoV-19/Wales/MILK-9A6A8D/2020\|EPI_ISL_550251\|2020 | ···················· | ···················· | ····················· | ···················· | ···················· | ···················· |
| J | 3344 | hCoV-19/England/MILK-9A8B1F/2020\|EPI_ISL_550252\|20 | ···················· | ···················· | ····················· | ···················· | ···················· | ···················· |
| J | 3345 | hCoV-19/England/MILK-9A52A4/2020\|EPI_ISL_550253\|20 | ···················· | ···················· | ····················· | ···················· | ···················· | ···················· |
| J | 3346 | hCoV-19/England/MILK-9A8335/2020\|EPI_ISL_550254\|20 | ···················· | ···················· | ····················· | ···················· | ···················· | ···················· |
| J | 3347 | hCoV-19/England/MILK-9A8450/2020\|EPI_ISL_550255\|20 | ···················· | ···················· | ····················· | ···················· | ···················· | ···················· |
| J | 3348 | hCoV-19/England/MILK-9A565D/2020\|EPI_ISL_550256\|20 | ···················· | ···················· | ····················· | ···················· | ···················· | ···················· |
| J | 3349 | hCoV-19/Wales/MILK-9A5338/2020\|EPI_ISL_550257\|2020 | ···················· | ···················· | ····················· | ···················· | ···················· | ···················· |
| J | 3350 | hCoV-19/England/MILK-9A8C3A/2020\|EPI_ISL_550258\|20 | ···················· | ···················· | ····················· | ···················· | ···················· | ···················· |
| J | 3351 | hCoV-19/England/MILK-9A85A8/2020\|EPI_ISL_550259\|20 | ···················· | ···················· | ····················· | ···················· | ···················· | ···················· |
| J | 3352 | hCoV-19/England/MILK-9A6980/2020\|EPI_ISL_550260\|20 | ···················· | ···················· | ·········T··········· | ···················· | ······T············· | ···················· |
| J | 3353 | hCoV-19/England/MILK-99A284/2020\|EPI_ISL_550262\|20 | ···················· | ···················· | ····················· | ···················· | ···················· | ···················· |
| J | 3354 | hCoV-19/England/MILK-99A80A/2020\|EPI_ISL_550263\|20 | ···················· | ···················· | ····················· | ···················· | ···················· | ···················· |
| J | 3355 | hCoV-19/England/MILK-999F33/2020\|EPI_ISL_550264\|20 | ···················· | ···················· | ····················· | ···················· | ···················· | ···················· |
| J | 3356 | hCoV-19/England/MILK-99A15A/2020\|EPI_ISL_550265\|20 | ···················· | ···················· | ····················· | ···················· | ···················· | ···················· |
| J | 3357 | hCoV-19/England/MILK-99A363/2020\|EPI_ISL_550273\|20 | ···················· | ···················· | ····················· | ···················· | ···················· | ···················· |
| J | 3358 | hCoV-19/England/MILK-99A451/2020\|EPI_ISL_550274\|20 | ···················· | ·A·················· | ····················· | ···················· | ···················· | ··A················· |
| J | 3359 | hCoV-19/England/MILK-99A002/2020\|EPI_ISL_550275\|20 | ···················· | ···················· | ····················· | ···················· | ···················· | ···················· |
| J | 3360 | hCoV-19/England/MILK-994439/2020\|EPI_ISL_551578\|20 | ···················· | ···················· | ····················· | ···················· | ···················· | ···················· |
| J | 3361 | hCoV-19/England/MILK-995562/2020\|EPI_ISL_551579\|20 | ···················· | ···················· | ····················· | ···················· | ···················· | ···················· |
| J | 3362 | hCoV-19/England/MILK-9940F9/2020\|EPI_ISL_551581\|20 | ···················· | ·A·················· | ····················· | ···················· | ···················· | ··A················· |
| J | 3363 | hCoV-19/England/MILK-995720/2020\|EPI_ISL_551582\|20 | ···················· | ···················· | ····················· | ···················· | ···················· | ···················· |
| J | 3364 | hCoV-19/England/MILK-99567E/2020\|EPI_ISL_551583\|20 | ···················· | ···················· | ····················· | ···················· | ···················· | ···················· |
| J | 3365 | hCoV-19/Wales/MILK-995B42/2020\|EPI_ISL_551584\|2020 | ···················· | ···················· | ····················· | ···················· | ···················· | ···················· |
| J | 3366 | hCoV-19/Wales/MILK-993F1B/2020\|EPI_ISL_551585\|2020 | ···················· | ···················· | ····················· | ···················· | ···················· | ···················· |
| J | 3367 | hCoV-19/England/MILK-993FFD/2020\|EPI_ISL_551587\|20 | ···················· | ···················· | ····················· | ···················· | ···················· | ···················· |
| J | 3368 | hCoV-19/England/MILK-995650/2020\|EPI_ISL_551588\|20 | ···················· | ···················· | ····················· | ···················· | ···················· | ···················· |
| J | 3369 | hCoV-19/England/MILK-994035/2020\|EPI_ISL_551589\|20 | ···················· | ···················· | ····················· | ···················· | ···················· | ···················· |
| J | 3370 | hCoV-19/England/MILK-9956F6/2020\|EPI_ISL_551590\|20 | ···················· | ···················· | ····················· | ···················· | ···················· | ···················· |
| J | 3371 | hCoV-19/England/MILK-9957C6/2020\|EPI_ISL_551591\|20 | ···················· | ···················· | ····················· | ···················· | ···················· | ···················· |
| J | 3372 | hCoV-19/England/MILK-995AAF/2020\|EPI_ISL_551592\|20 | ···················· | ·A·················· | ····················· | ···················· | ···················· | ···················· |
| J | 3373 | hCoV-19/England/MILK-99584B/2020\|EPI_ISL_551594\|20 | ···················· | ···················· | ····················· | ···················· | ···················· | ···················· |
| J | 3374 | hCoV-19/England/MILK-99583C/2020\|EPI_ISL_551595\|20 | ···················· | ···················· | ····················· | ···················· | ···················· | ···················· |
| J | 3375 | hCoV-19/England/MILK-994071/2020\|EPI_ISL_551596\|20 | ···················· | ···················· | ····················· | ···················· | ···················· | ···················· |
| J | 3376 | hCoV-19/England/ALDP-925A2C/2020\|EPI_ISL_552801\|20 | ···················· | ···················· | ····················· | ···················· | ···················· | ···················· |
| J | 3377 | hCoV-19/England/MILK-93A462/2020\|EPI_ISL_552802\|20 | ···················· | ···················· | ····················· | ···················· | ···················· | ···················· |
| J | 3378 | hCoV-19/England/ALDP-928977/2020\|EPI_ISL_552803\|20 | ···················· | ···················· | ····················· | ···················· | ···················· | ···················· |
| J | 3379 | hCoV-19/England/ALDP-929026/2020\|EPI_ISL_552804\|20 | ···················· | ···················· | ····················· | ···················· | ···················· | ···················· |
| J | 3380 | hCoV-19/England/ALDP-920BB5/2020\|EPI_ISL_552805\|20 | ···················· | ·A·················· | ····················· | ···················· | ···················· | ···················· |
| J | 3381 | hCoV-19/England/ALDP-93138C/2020\|EPI_ISL_552806\|20 | ···················· | ·A·················· | ····················· | ···················· | ···················· | ···················· |
| J | 3382 | hCoV-19/England/ALDP-92992B/2020\|EPI_ISL_552807\|20 | ···················· | ···················· | ····················· | ···················· | ···················· | ···················· |
| J | 3383 | hCoV-19/England/ALDP-926F1D/2020\|EPI_ISL_552808\|20 | ···················· | ···················· | ····················· | ···················· | ···················· | ···················· |
| J | 3384 | hCoV-19/England/MILK-93C9BC/2020\|EPI_ISL_552809\|20 | ···················· | ···················· | ····················· | ···················· | ···················· | ···················· |
| J | 3385 | hCoV-19/England/MILK-93E8F9/2020\|EPI_ISL_552810\|20 | ···················· | ···················· | ····················· | ···················· | ···················· | ···················· |
| J | 3386 | hCoV-19/England/MILK-93CFE7/2020\|EPI_ISL_552811\|20 | ···················· | ···················· | ····················· | ···················· | ···················· | ···················· |
| J | 3387 | hCoV-19/England/ALDP-92686D/2020\|EPI_ISL_552812\|20 | ···················· | ···················· | ····················· | ···················· | ···················· | ···················· |
| J | 3388 | hCoV-19/England/ALDP-929D4D/2020\|EPI_ISL_552813\|20 | ···················· | ···················· | ····················· | ···················· | ···················· | ···················· |
| J | 3389 | hCoV-19/England/MILK-93EBD2/2020\|EPI_ISL_552814\|20 | ···················· | ···················· | ····················· | ···················· | ···················· | ···················· |
| J | 3390 | hCoV-19/England/ALDP-9353B5/2020\|EPI_ISL_552815\|20 | ···················· | ·A·················· | ····················· | ···················· | ···················· | ···················· |
| J | 3391 | hCoV-19/England/ALDP-9241BF/2020\|EPI_ISL_552817\|20 | ···················· | ···················· | ····················· | ···················· | ···················· | ···················· |
| J | 3392 | hCoV-19/England/ALDP-95AC82/2020\|EPI_ISL_555001\|20 | ···················· | ···················· | ····················· | ···················· | ···················· | ···················· |
| J | 3393 | hCoV-19/England/ALDP-94D7CD/2020\|EPI_ISL_555002\|20 | ···················· | ···················· | ····················· | ···················· | ···················· | ···················· |
| J | 3394 | hCoV-19/England/ALDP-95CE5D/2020\|EPI_ISL_555004\|20 | ···················· | ···················· | ····················· | ···················· | ···················· | ···················· |
| J | 3395 | hCoV-19/Wales/ALDP-95AB2B/2020\|EPI_ISL_555005\|2020 | ···················· | ···················· | ····················· | ···················· | ···················· | ···················· |
| J | 3396 | hCoV-19/England/ALDP-94D60C/2020\|EPI_ISL_555006\|20 | ···················· | ···················· | ····················· | ···················· | ···················· | ···················· |
| J | 3397 | hCoV-19/England/ALDP-95C752/2020\|EPI_ISL_555007\|20 | ···················· | ···················· | ····················· | ···················· | ···················· | ···················· |
| J | 3398 | hCoV-19/England/ALDP-95AC55/2020\|EPI_ISL_555008\|20 | ···················· | ···················· | ····················· | ···················· | ···················· | ···················· |
| J | 3399 | hCoV-19/England/ALDP-95AA79/2020\|EPI_ISL_555009\|20 | ···················· | ···················· | ····················· | ···················· | ···················· | ···················· |
| J | 3400 | hCoV-19/England/ALDP-95C725/2020\|EPI_ISL_555013\|20 | ···················· | ···················· | ····················· | ···················· | ···················· | ···················· |
| J | 3401 | hCoV-19/England/ALDP-94D6DF/2020\|EPI_ISL_555014\|20 | ···················· | ···················· | ····················· | ···················· | ···················· | ···················· |
| J | 3402 | hCoV-19/England/ALDP-95CD23/2020\|EPI_ISL_555015\|20 | ···················· | ···················· | ····················· | ···················· | ···················· | ···················· |
| J | 3403 | hCoV-19/England/ALDP-94D596/2020\|EPI_ISL_555017\|20 | ···················· | ···················· | ····················· | ···················· | ···················· | ···················· |
| J | 3404 | hCoV-19/England/ALDP-95C0DF/2020\|EPI_ISL_555071\|20 | ···················· | ···················· | ····················· | ···················· | ···················· | ···················· |
| J | 3405 | hCoV-19/England/ALDP-95A639/2020\|EPI_ISL_555072\|20 | ···················· | ···················· | ····················· | ···················· | ···················· | ···················· |
| J | 3406 | hCoV-19/England/ALDP-95C2F7/2020\|EPI_ISL_555074\|20 | ···················· | ···················· | ····················· | ···················· | ···················· | ···················· |
| J | 3407 | hCoV-19/England/ALDP-95C628/2020\|EPI_ISL_555075\|20 | ···················· | ···················· | ····················· | ···················· | ···················· | ···················· |
| J | 3408 | hCoV-19/England/ALDP-95C3A9/2020\|EPI_ISL_555076\|20 | ···················· | ···················· | ····················· | ···················· | ···················· | ···················· |
| J | 3409 | hCoV-19/England/ALDP-95C251/2020\|EPI_ISL_555077\|20 | ···················· | ···················· | ····················· | ···················· | ···················· | ···················· |
| J | 3410 | hCoV-19/England/ALDP-94D55A/2020\|EPI_ISL_555079\|20 | ···················· | ···················· | ····················· | ···················· | ···················· | ···················· |
| J | 3411 | hCoV-19/England/ALDP-95C7E9/2020\|EPI_ISL_555081\|20 | ···················· | ···················· | ····················· | ···················· | ···················· | ···················· |
| J | 3412 | hCoV-19/England/ALDP-95A790/2020\|EPI_ISL_555082\|20 | ···················· | ···················· | ····················· | ···················· | ···················· | ···················· |
| J | 3413 | hCoV-19/England/ALDP-94E96C/2020\|EPI_ISL_555083\|20 | ···················· | ···················· | ····················· | ···················· | ···················· | ···················· |
| J | 3414 | hCoV-19/England/ALDP-94EA96/2020\|EPI_ISL_555085\|20 | ···················· | ···················· | ····················· | ···················· | ···················· | ···················· |
| J | 3415 | hCoV-19/England/ALDP-95C6A0/2020\|EPI_ISL_555086\|20 | ···················· | ·············A······ | ····················· | ···················· | ···················· | ···················· |
| J | 3416 | hCoV-19/England/ALDP-95C52B/2020\|EPI_ISL_555087\|20 | ···················· | ···················· | ····················· | ···················· | ···················· | ···················· |
| J | 3417 | hCoV-19/England/ALDP-95CFF0/2020\|EPI_ISL_555088\|20 | ···················· | ···················· | ····················· | ···················· | ···················· | ···················· |

J 3418 hCoV-19/England/ALDP-959D53/2020|EPI_ISL_555089|20 ···················· ···················· ····················· ···················· ···················· ····················
J 3419 hCoV-19/England/ALDP-95C2D9/2020|EPI_ISL_555090|20 ···················· ···················· ····················· ···················· ···················· ····················
J 3420 hCoV-19/England/ALDP-95CD6F/2020|EPI_ISL_555092|20 ···················· ···················· ····················· ···················· ···················· ····················
J 3421 hCoV-19/England/ALDP-95CF87/2020|EPI_ISL_555093|20 ···················· ·············A······ ····················· ···················· ···················· ····················
J 3422 hCoV-19/England/ALDP-94D684/2020|EPI_ISL_555094|20 ···················· ···················· ····················· ···················· ···················· ····················
J 3423 hCoV-19/England/ALDP-95C093/2020|EPI_ISL_555095|20 ···················· ·············A······ ····················· ···················· ···················· ····················
J 3424 hCoV-19/England/ALDP-94D6FD/2020|EPI_ISL_555098|20 ···················· ···················· ····················· ···················· ···················· ····················
J 3425 hCoV-19/England/ALDP-94D61B/2020|EPI_ISL_555099|20 ···················· ···················· ····················· ···················· ···················· ····················
J 3426 hCoV-19/England/MILK-6303DB/2020|EPI_ISL_556300|20 ···················· ·A·················· ····················· ···················· ···················· ···A················
J 3427 hCoV-19/England/ALDP-638A56/2020|EPI_ISL_556301|20 ···················· ···················· ····················· ···················· ···················· ····················
J 3428 hCoV-19/England/ALDP-6CFA54/2020|EPI_ISL_556302|20 ···················· ···················· ····················· ···················· ···················· ····················
J 3429 hCoV-19/England/ALDP-6D46D0/2020|EPI_ISL_556303|20 ···················· ···················· ····················· ···················· ···················· ····················
J 3430 hCoV-19/Wales/MILK-95DA85/2020|EPI_ISL_556304|2020 ···················· ···················· ····················· ···················· ···················· ····················
J 3431 hCoV-19/England/ALDP-94AF7D/2020|EPI_ISL_556306|20 ···················· ···················· ····················· ···················· ···················· ····················
J 3432 hCoV-19/England/ALDP-94D2F9/2020|EPI_ISL_556307|20 ···················· ···················· ····················· ···················· ···················· ····················
J 3433 hCoV-19/England/CAMC-94B5E3/2020|EPI_ISL_556309|20 ···················· ···················· ····················· ···················· ···················· ····················
J 3434 hCoV-19/England/ALDP-949C2C/2020|EPI_ISL_556310|20 ···················· ···················· ····················· ···················· ···················· ····················
J 3435 hCoV-19/England/ALDP-94A186/2020|EPI_ISL_556311|20 ···················· ···················· ····················· ···················· ···················· ····················
J 3436 hCoV-19/England/ALDP-94D305/2020|EPI_ISL_556312|20 ···················· ···················· ····················· ···················· ···················· ····················
J 3437 hCoV-19/England/ALDP-94A7DF/2020|EPI_ISL_556314|20 ···················· ···················· ····················· ···················· ···················· ····················
J 3438 hCoV-19/England/ALDP-94D37E/2020|EPI_ISL_556315|20 ···················· ···················· ····················· ···················· ···················· ····················
J 3439 hCoV-19/England/CAMC-94B4E6/2020|EPI_ISL_556318|20 ···················· ···················· ····················· ···················· ···················· ····················
J 3440 hCoV-19/England/ALDP-94D3AB/2020|EPI_ISL_556319|20 ···················· ·······K············ ····················· ···················· ···················· ····················
J 3441 hCoV-19/England/ALDP-94D2DB/2020|EPI_ISL_556320|20 ···················· ···················· ····················· ···················· ···················· ····················
J 3442 hCoV-19/England/CAMC-949CFF/2020|EPI_ISL_556321|20 ···················· ···················· ····················· ···················· ···················· ····················
J 3443 hCoV-19/England/ALDP-94A845/2020|EPI_ISL_556323|20 ···················· ···················· ····················· ···················· ···················· ····················
J 3444 hCoV-19/England/ALDP-94D323/2020|EPI_ISL_556325|20 ···················· ···················· ····················· ···················· ···················· ····················
J 3445 hCoV-19/England/ALDP-94ED33/2020|EPI_ISL_556326|20 ···················· ···················· ····················· ···················· ···················· ····················
J 3446 hCoV-19/England/ALDP-94A05C/2020|EPI_ISL_556327|20 ···················· ···················· ····················· ···················· ···················· ····················
J 3447 hCoV-19/England/ALDP-94F1AF/2020|EPI_ISL_556328|20 ···················· ·A·················· ····················· ···················· ···················· ····················
J 3448 hCoV-19/England/ALDP-94ED7F/2020|EPI_ISL_556329|20 ···················· ···················· ····················· ···················· ···················· ····················
J 3449 hCoV-19/England/ALDP-949B2F/2020|EPI_ISL_556331|20 ···················· ···················· ····················· ···················· ···················· ····················
J 3450 hCoV-19/England/ALDP-94EFD3/2020|EPI_ISL_556332|20 ···················· ···················· ····················· ···················· ···················· ····················
J 3451 hCoV-19/England/ALDP-94B6A4/2020|EPI_ISL_556333|20 ···················· ···················· ····················· ···················· ···················· ····················
J 3452 hCoV-19/England/ALDP-94D1A1/2020|EPI_ISL_556334|20 ···················· ·A·················· ····················· ···················· ···················· ····················
J 3453 hCoV-19/England/ALDP-9487B3/2020|EPI_ISL_556335|20 ···················· ···················· ····················· ···················· ···················· ····················
J 3454 hCoV-19/England/ALDP-94A6F0/2020|EPI_ISL_556336|20 ···················· ···················· ····················· ···················· ···················· ····················
J 3455 hCoV-19/England/ALDP-94EE03/2020|EPI_ISL_556337|20 ···················· ···················· ····················· ···················· ···················· ····················
J 3456 hCoV-19/England/CAMC-94B6FF/2020|EPI_ISL_556338|20 ···················· ···················· ····················· ···················· ···················· ····················
J 3457 hCoV-19/England/ALDP-949A22/2020|EPI_ISL_556339|20 ···················· ···················· ····················· ···················· ···················· ····················
J 3458 hCoV-19/England/ALDP-94D45D/2020|EPI_ISL_556341|20 ···················· ···················· ····················· ···················· ···················· ····················
J 3459 hCoV-19/England/ALDP-94D43F/2020|EPI_ISL_556342|20 ···················· ···················· ····················· ···················· ···················· ····················
J 3460 hCoV-19/England/ALDP-94B905/2020|EPI_ISL_556343|20 ···················· ···················· ····················· ···················· ···················· ····················
J 3461 hCoV-19/England/CAMC-94B70B/2020|EPI_ISL_556344|20 ···················· ···················· ····················· ···················· ···················· ····················
J 3462 hCoV-19/England/ALDP-94AF13/2020|EPI_ISL_556345|20 ···················· ···················· ····················· ···················· ···················· ····················
J 3463 hCoV-19/England/ALDP-94D1B0/2020|EPI_ISL_556346|20 ···················· ·A·················· ····················· ···················· ···················· ····················
J 3464 hCoV-19/England/ALDP-94D208/2020|EPI_ISL_556347|20 ···················· ···················· ····················· ···················· ···················· ····················
J 3465 hCoV-19/England/ALDP-6E9C4F/2020|EPI_ISL_557600|20 ···················· ···················· ····················· ···················· ···················· ····················
J 3466 hCoV-19/England/ALDP-70814A/2020|EPI_ISL_557601|20 ···················· ·A·················· ····················· ···················· ···················· ····················
J 3467 hCoV-19/England/ALDP-70720C/2020|EPI_ISL_557602|20 ···················· ·A·················· ····················· ···················· ···················· ····················
J 3468 hCoV-19/England/ALDP-71E0EA/2020|EPI_ISL_557603|20 ···················· ···················· ····················· ···················· ···················· ····················
J 3469 hCoV-19/England/ALDP-71CE1F/2020|EPI_ISL_557605|20 ···················· ·A·················· ····················· ···················· ···················· ····················
J 3470 hCoV-19/England/ALDP-706CC3/2020|EPI_ISL_557606|20 ···················· ···················· ····················· ···················· ···················· ····················
J 3471 hCoV-19/England/ALDP-6E97E4/2020|EPI_ISL_557608|20 ···················· ···················· ····················· ···················· ···················· ····················
J 3472 hCoV-19/England/ALDP-6E5B82/2020|EPI_ISL_557609|20 ···················· ···················· ····················· ···················· ····T··············· ····················
J 3473 hCoV-19/England/ALDP-6EB849/2020|EPI_ISL_557610|20 ···················· ·A·················· ····················· ···················· ···················· ····················
J 3474 hCoV-19/England/ALDP-6E8994/2020|EPI_ISL_557611|20 ···················· ···················· ····················· ···················· ···················· ····················
J 3475 hCoV-19/England/ALDP-7054AE/2020|EPI_ISL_557613|20 ···················· ·A·················· ····················· ···················· ···················· ····················
J 3476 hCoV-19/England/ALDP-6EAB32/2020|EPI_ISL_557614|20 ···················· ···················· ····················· ···················· ···················· ····················
J 3477 hCoV-19/England/ALDP-703534/2020|EPI_ISL_557616|20 ···················· ···················· ····················· ···················· ···················· ····················
J 3478 hCoV-19/England/ALDP-6EA7B6/2020|EPI_ISL_557617|20 ···················· ···················· ····················· ···················· ···················· ····················
J 3479 hCoV-19/England/ALDP-6E14F4/2020|EPI_ISL_557618|20 ···················· ···················· ····················· ···················· ···················· ····················
J 3480 hCoV-19/England/ALDP-705013/2020|EPI_ISL_557619|20 ···················· ···················· ····················· ···················· ···················· ····················
J 3481 hCoV-19/England/ALDP-71CF0D/2020|EPI_ISL_557620|20 ···················· ···················· ····················· ···················· ···················· ····················
J 3482 hCoV-19/England/ALDP-6E2806/2020|EPI_ISL_557621|20 ···················· ···················· ····················· ···················· ···················· ····················
J 3483 hCoV-19/England/ALDP-6E9ADC/2020|EPI_ISL_557622|20 ···················· ···················· ····················· ···················· ···················· ····················
J 3484 hCoV-19/England/ALDP-722D28/2020|EPI_ISL_557623|20 ···················· ···················· ····················· ···················· ···················· ····················
J 3485 hCoV-19/England/ALDP-720EBE/2020|EPI_ISL_557624|20 ···················· ···················· ····················· ···················· ···················· ····················
J 3486 hCoV-19/England/ALDP-71EDC5/2020|EPI_ISL_557627|20 ···················· ···················· ····················· ···················· ···················· ····················
J 3487 hCoV-19/England/ALDP-6E9D88/2020|EPI_ISL_557628|20 ···················· ···················· ····················· ···················· ···················· ····················
J 3488 hCoV-19/England/ALDP-704A61/2020|EPI_ISL_557631|20 ···················· ···················· ····················· ···················· ···················· ····················
J 3489 hCoV-19/England/ALDP-6E1B68/2020|EPI_ISL_557632|20 ···················· ···················· ····················· ···················· ···················· ····················
J 3490 hCoV-19/England/ALDP-6DE16C/2020|EPI_ISL_557633|20 ···················· ···················· ····················· ···················· ···················· ····················
J 3491 hCoV-19/England/ALDP-6EA9A1/2020|EPI_ISL_557634|20 ···················· ···················· ····················· ···················· ···················· ····················
J 3492 hCoV-19/England/ALDP-6E8F0B/2020|EPI_ISL_557635|20 ···················· ···················· ····················· ···················· ···················· ····················
J 3493 hCoV-19/England/ALDP-6E51BD/2020|EPI_ISL_557636|20 ···················· ···················· ····················· ···················· ···················· ····················
J 3494 hCoV-19/England/ALDP-6E8F38/2020|EPI_ISL_557637|20 ···················· ···················· ····················· ···················· ···················· ····················
J 3495 hCoV-19/England/ALDP-724A89/2020|EPI_ISL_557638|20 ···················· ···················· ····················· ···················· ···················· ····················
J 3496 hCoV-19/England/ALDP-71DF66/2020|EPI_ISL_557639|20 ···················· ···················· ····················· ···················· ···················· ····················
J 3497 hCoV-19/England/ALDP-720CD2/2020|EPI_ISL_557640|20 ···················· ···················· ····················· ···················· ···················· ····················

```
J 3498 hCoV-19/England/ALDP-71D2C7/2020|EPI_ISL_557641|20 ···················· ···················· ····················· ···················· ···················· ····················
J 3499 hCoV-19/England/ALDP-71D151/2020|EPI_ISL_557642|20 ···················· ···················· ····················· ···················· ···················· ····················
J 3500 hCoV-19/England/ALDP-6DF462/2020|EPI_ISL_557644|20 ···················· ···················· ····················· ···················· ···················· ····················
J 3501 hCoV-19/England/ALDP-6E492F/2020|EPI_ISL_557645|20 ···················· ···················· ····················· ···················· ···················· ····················
J 3502 hCoV-19/England/ALDP-71E378/2020|EPI_ISL_557646|20 ···················· ···················· ····················· ···················· ···················· ····················
J 3503 hCoV-19/England/ALDP-6EB050/2020|EPI_ISL_557648|20 ···················· ···················· ····················· ···················· ···················· ····················
J 3504 hCoV-19/England/ALDP-71DF93/2020|EPI_ISL_557649|20 ···················· ···················· ····················· ···················· ···················· ····················
J 3505 hCoV-19/England/ALDP-71C4A4/2020|EPI_ISL_557650|20 ···················· ···················· ····················· ···················· ···················· ····················
J 3506 hCoV-19/England/ALDP-720C1E/2020|EPI_ISL_557651|20 ···················· ···················· ····················· ···················· ···················· ····················
J 3507 hCoV-19/England/ALDP-71C019/2020|EPI_ISL_557652|20 ···················· ···················· ····················· ···················· ···················· ····················
J 3508 hCoV-19/England/ALDP-720BF3/2020|EPI_ISL_557653|20 ···················· ···················· ····················· ···················· ···················· ····················
J 3509 hCoV-19/England/ALDP-71C51A/2020|EPI_ISL_557654|20 ···················· ···················· ····················· ···················· ···················· ····················
J 3510 hCoV-19/England/ALDP-70D39A/2020|EPI_ISL_557656|20 ···················· ···················· ····················· ···················· ···················· ····················
J 3511 hCoV-19/England/ALDP-6E26C0/2020|EPI_ISL_557657|20 ···················· ···················· ····················· ···················· ···················· ····················
J 3512 hCoV-19/England/ALDP-7214CA/2020|EPI_ISL_557659|20 ···················· ···················· ····················· ···················· ···················· ····················
J 3513 hCoV-19/England/ALDP-6DDFDC/2020|EPI_ISL_557661|20 ········T··········· ···················· ····················· ···················· ···················· ····················
J 3514 hCoV-19/England/ALDP-709905/2020|EPI_ISL_557662|20 ···················· ···················· ····················· ···················· ···················· ····················
J 3515 hCoV-19/England/ALDP-71D160/2020|EPI_ISL_557663|20 ···················· ···················· ····················· ···················· ···················· ····················
J 3516 hCoV-19/England/ALDP-709F21/2020|EPI_ISL_557664|20 ···················· ···················· ····················· ···················· ···················· ····················
J 3517 hCoV-19/England/ALDP-6E21FC/2020|EPI_ISL_557665|20 ···················· ···················· ····················· ···················· ···················· ····················
J 3518 hCoV-19/England/ALDP-7259C7/2020|EPI_ISL_557666|20 ···················· ···················· ····················· ···················· ···················· ····················
J 3519 hCoV-19/England/ALDP-6EB80D/2020|EPI_ISL_557667|20 ···················· ···················· ····················· ···················· ···················· ····················
J 3520 hCoV-19/England/ALDP-6E90BC/2020|EPI_ISL_557668|20 ···················· ···················· ····················· ···················· ···················· ····················
J 3521 hCoV-19/England/ALDP-704CF2/2020|EPI_ISL_557669|20 ···················· ···················· ····················· ···················· ···················· ····················
J 3522 hCoV-19/England/ALDP-6E0510/2020|EPI_ISL_557670|20 ···················· ···················· ····················· ···················· ···················· ····················
J 3523 hCoV-19/England/ALDP-6E95EA/2020|EPI_ISL_557671|20 ···················· ···················· ····················· ···················· ···················· ····················
J 3524 hCoV-19/England/ALDP-7210F3/2020|EPI_ISL_557673|20 ···················· ···················· ····················· ···················· ···················· ····················
J 3525 hCoV-19/England/ALDP-70B4A8/2020|EPI_ISL_557674|20 ···················· ···················· ····················· ···················· ···················· ····················
J 3526 hCoV-19/England/ALDP-721196/2020|EPI_ISL_557677|20 ···················· ···················· ····················· ···················· ···················· ····················
J 3527 hCoV-19/England/ALDP-6DF9EB/2020|EPI_ISL_557678|20 ···················· ···················· ····················· ···················· ···················· ····················
J 3528 hCoV-19/England/ALDP-6FF806/2020|EPI_ISL_557679|20 ···················· ·A·················· ····················· ···················· ···················· ····················
J 3529 hCoV-19/England/MILK-51856B/2020|EPI_ISL_558923|20 ···················· ···················· ····················· ···················· ···················· ····················
J 3530 hCoV-19/England/ALDP-52A89A/2020|EPI_ISL_558924|20 ···················· ···················· ····················· ···················· ···················· ····················
J 3531 hCoV-19/England/MILK-5149AF/2020|EPI_ISL_558925|20 ···················· ···················· ····················· ···················· ···················· ····················
J 3532 hCoV-19/England/ALDP-526421/2020|EPI_ISL_558927|20 ···················· ···················· ····················· ···················· ···················· ····················
J 3533 hCoV-19/England/MILK-50ED60/2020|EPI_ISL_558928|20 ···················· ···················· ····················· ···················· ···················· ····················
J 3534 hCoV-19/England/ALDP-52B8A8/2020|EPI_ISL_558929|20 ···················· ···················· ····················· ···················· ···················· ····················
J 3535 hCoV-19/England/ALDP-51D751/2020|EPI_ISL_558930|20 ···················· ···················· ····················· ···················· ···················· ····················
J 3536 hCoV-19/England/ALDP-52AF3B/2020|EPI_ISL_558931|20 ···················· ···················· ····················· ···················· ···················· ····················
J 3537 hCoV-19/England/MILK-5149CD/2020|EPI_ISL_558932|20 ···················· ···················· ····················· ···················· ···················· ····················
J 3538 hCoV-19/England/ALDP-52AB46/2020|EPI_ISL_558933|20 ···················· ···················· ····················· ···················· ···················· ····················
J 3539 hCoV-19/England/MILK-50C227/2020|EPI_ISL_558934|20 ···················· ···················· ····················· ···················· ···················· ····················
J 3540 hCoV-19/England/ALDP-522935/2020|EPI_ISL_558936|20 ···················· ···················· ····················· ···················· ···················· ····················
J 3541 hCoV-19/England/ALDP-522328/2020|EPI_ISL_558938|20 ···················· ···················· ····················· ···················· ···················· ····················
J 3542 hCoV-19/England/ALDP-5290B1/2020|EPI_ISL_558939|20 ···················· ···················· ····················· ···················· ···················· ····················
J 3543 hCoV-19/England/ALDP-523C2C/2020|EPI_ISL_558940|20 ···················· ···················· ····················· ···················· ···················· ····················
J 3544 hCoV-19/England/ALDP-51D5B1/2020|EPI_ISL_558941|20 ···················· ···················· ····················· ···················· ···················· ····················
J 3545 hCoV-19/England/ALDP-524247/2020|EPI_ISL_558942|20 ···················· ···················· ····················· ···················· ···················· ····················
J 3546 hCoV-19/England/ALDP-51F28B/2020|EPI_ISL_558944|20 ···················· ···················· ····················· ···················· ···················· ····················
J 3547 hCoV-19/England/ALDP-521532/2020|EPI_ISL_558945|20 ···················· ···················· ····················· ···················· ···················· ····················
J 3548 hCoV-19/England/ALDP-51FEF0/2020|EPI_ISL_558948|20 ···················· ···················· ····················· ···················· ···················· ····················
J 3549 hCoV-19/England/MILK-50FEF3/2020|EPI_ISL_558951|20 ···················· ···················· ····················· ···················· ···················· ····················
J 3550 hCoV-19/England/ALDP-51D8E5/2020|EPI_ISL_558952|20 ···················· ···················· ····················· ···················· ···················· ····················
J 3551 hCoV-19/England/ALDP-5218A2/2020|EPI_ISL_558954|20 ···················· ···················· ····················· ···················· ···················· ····················
J 3552 hCoV-19/England/ALDP-5240D4/2020|EPI_ISL_558956|20 ···················· ···················· ····················· ···················· ···················· ····················
J 3553 hCoV-19/England/ALDP-52ADC8/2020|EPI_ISL_558957|20 ···················· ···················· ····················· ···················· ···················· ····················
J 3554 hCoV-19/England/ALDP-52C40D/2020|EPI_ISL_558958|20 ···················· ···················· ···········T········· ···················· ···················· ····················
J 3555 hCoV-19/England/MILK-50CB95/2020|EPI_ISL_558959|20 ···················· ·A·················· ····················· ···················· ···················· ····················
J 3556 hCoV-19/England/ALDP-51D645/2020|EPI_ISL_558960|20 ···················· ···················· ····················· ···················· ···················· ····················
J 3557 hCoV-19/England/ALDP-519DCC/2020|EPI_ISL_558961|20 ···················· ···················· ····················· ···················· ···················· ····················
J 3558 hCoV-19/England/ALDP-5292F7/2020|EPI_ISL_558962|20 ···················· ···················· ····················· ···················· ···················· ····················
J 3559 hCoV-19/England/ALDP-526C56/2020|EPI_ISL_558963|20 ···················· ···················· ····················· ···················· ···················· ····················
J 3560 hCoV-19/England/ALDP-526FC6/2020|EPI_ISL_558964|20 ···················· ···················· ····················· ···················· ···················· ····················
J 3561 hCoV-19/England/MILK-50E38C/2020|EPI_ISL_558965|20 ···················· ···················· ····················· ···················· ···················· ····················
J 3562 hCoV-19/England/ALDP-52A6AF/2020|EPI_ISL_558966|20 ···················· ···················· ····················· ···················· ···················· ····················
J 3563 hCoV-19/England/ALDP-52B88A/2020|EPI_ISL_558967|20 ···················· ·A·················· ····················· ···················· ···················· ····················
J 3564 hCoV-19/England/ALDP-5244C9/2020|EPI_ISL_558968|20 ···················· ···················· ····················· ···················· ···················· ····················
J 3565 hCoV-19/England/ALDP-52B1DA/2020|EPI_ISL_558969|20 ···················· ···················· ····················· ···················· ···················· ····················
J 3566 hCoV-19/England/MILK-515012/2020|EPI_ISL_558970|20 ···················· ···················· ····················· ···················· ···················· ····················
J 3567 hCoV-19/England/ALDP-52B87B/2020|EPI_ISL_558971|20 ···················· ···················· ····················· ···················· ···················· ····················
J 3568 hCoV-19/England/ALDP-5217D2/2020|EPI_ISL_558972|20 ···················· ···················· ···········T········· ···················· ···················· ····················
J 3569 hCoV-19/England/MILK-5169F8/2020|EPI_ISL_558973|20 ···················· ···················· ····················· ···················· ···················· ····················
J 3570 hCoV-19/England/ALDP-51DEF2/2020|EPI_ISL_558976|20 ···················· ···················· ····················· ···················· ···················· ····················
J 3571 hCoV-19/England/ALDP-524C76/2020|EPI_ISL_558979|20 ···················· ···················· ····················· ···················· ···················· ····················
J 3572 hCoV-19/England/MILK-50EF97/2020|EPI_ISL_558980|20 ···················· ···················· ····················· ···················· ···················· ····················
J 3573 hCoV-19/England/MILK-50CF11/2020|EPI_ISL_558981|20 ···················· ···················· ····················· ···················· ···················· ····················
J 3574 hCoV-19/England/ALDP-524D73/2020|EPI_ISL_558982|20 ···················· ···················· ····················· ···················· ···················· ····················
J 3575 hCoV-19/England/ALDP-52ABA0/2020|EPI_ISL_558983|20 ···················· ···················· ····················· ···················· ···················· ····················
J 3576 hCoV-19/England/ALDP-525E9D/2020|EPI_ISL_558984|20 ···················· ···················· ····················· ···················· ···················· ····················
J 3577 hCoV-19/England/ALDP-529066/2020|EPI_ISL_558986|20 ···················· ···················· ····················· ···················· ···················· ····················
```

| | | | | | | | | |
|---|---|---|---|---|---|---|---|---|
| J | 3578 | hCoV-19/England/ALDP-521F9E/2020\|EPI_ISL_558988\|20 | ···················· | ···················· | ····················· | ···················· | ···················· | ···················· |
| J | 3579 | hCoV-19/England/ALDP-528A87/2020\|EPI_ISL_558989\|20 | ···················· | ···················· | ····················· | ···················· | ···················· | ···················· |
| J | 3580 | hCoV-19/England/ALDP-526CB0/2020\|EPI_ISL_558991\|20 | ···················· | ···················· | ····················· | ···················· | ···················· | ···················· |
| J | 3581 | hCoV-19/England/ALDP-519719/2020\|EPI_ISL_558992\|20 | ···················· | ···················· | ····················· | ···················· | ···················· | ···················· |
| J | 3582 | hCoV-19/England/ALDP-52A8C7/2020\|EPI_ISL_558997\|20 | ···················· | ···················· | ····················· | ···················· | ···················· | ···················· |
| J | 3583 | hCoV-19/England/ALDP-5279F4/2020\|EPI_ISL_558998\|20 | ···················· | ···················· | ····················· | ···················· | ···················· | ···················· |
| J | 3584 | hCoV-19/England/ALDP-52B2C8/2020\|EPI_ISL_558999\|20 | ···················· | ···················· | ····················· | ···················· | ···················· | ···················· |
| J | 3585 | hCoV-19/USA/FL-BPHL-1418/2020\|EPI_ISL_549200\|2020- | ···················· | ···················· | ····················· | ···················· | ···················· | ···················· |
| J | 3586 | hCoV-19/USA/FL-BPHL-1419/2020\|EPI_ISL_549201\|2020- | ···················· | ···················· | ····················· | ···················· | ···················· | ···················· |
| J | 3587 | hCoV-19/USA/FL-BPHL-1420/2020\|EPI_ISL_549202\|2020- | ···················· | ···················· | ····················· | ···················· | ···················· | ···················· |
| J | 3588 | hCoV-19/USA/FL-BPHL-1421/2020\|EPI_ISL_549203\|2020- | ···················· | ···················· | ····················· | ···················· | ···················· | ···················· |
| J | 3589 | hCoV-19/USA/FL-BPHL-1422/2020\|EPI_ISL_549204\|2020- | ···················· | ···················· | ····················· | ···················· | ···················· | ···················· |
| J | 3590 | hCoV-19/USA/FL-BPHL-1423/2020\|EPI_ISL_549205\|2020- | ···················· | ···················· | ····················· | ···················· | ···················· | ···················· |
| J | 3591 | hCoV-19/USA/FL-BPHL-1424/2020\|EPI_ISL_549206\|2020- | ···················· | ···················· | ····················· | ···················· | ···················· | ···················· |
| J | 3592 | hCoV-19/USA/FL-BPHL-1425/2020\|EPI_ISL_549207\|2020- | ···················· | ···················· | ····················· | ···················· | ···················· | ···················· |
| J | 3593 | hCoV-19/USA/FL-BPHL-1426/2020\|EPI_ISL_549208\|2020- | ···················· | ···················· | ····················· | ···················· | ···················· | ···················· |
| J | 3594 | hCoV-19/USA/FL-BPHL-1428/2020\|EPI_ISL_549210\|2020- | ···················· | ···················· | ····················· | ···················· | ···················· | ···················· |
| J | 3596 | hCoV-19/USA/FL-BPHL-1430/2020\|EPI_ISL_549212\|2020- | ···················· | ·A·················· | ····················· | ···················· | ···················· | ···················· |
| J | 3597 | hCoV-19/USA/FL-BPHL-1431/2020\|EPI_ISL_549213\|2020- | ···················· | ···················· | ····················· | ···················· | ···················· | ···················· |
| J | 3598 | hCoV-19/USA/FL-BPHL-1432/2020\|EPI_ISL_549214\|2020- | ···················· | ···················· | ····················· | ···················· | ···················· | ···················· |
| J | 3599 | hCoV-19/USA/FL-BPHL-1433/2020\|EPI_ISL_549215\|2020- | ···················· | ···················· | ····················· | ···················· | ···················· | ···················· |
| J | 3600 | hCoV-19/USA/FL-BPHL-1434/2020\|EPI_ISL_549216\|2020- | ···················· | ···················· | ····················· | ···················· | ···················· | ···················· |
| J | 3601 | hCoV-19/USA/FL-BPHL-1435/2020\|EPI_ISL_549217\|2020- | ···················· | ···················· | ····················· | ···················· | ···················· | ···················· |
| J | 3602 | hCoV-19/USA/FL-BPHL-1436/2020\|EPI_ISL_549218\|2020- | ···················· | ···················· | ····················· | ···················· | ···················· | ···················· |
| J | 3603 | hCoV-19/USA/FL-BPHL-1437/2020\|EPI_ISL_549219\|2020- | ···················· | ···················· | ····················· | ···················· | ···················· | ···················· |
| J | 3604 | hCoV-19/USA/FL-BPHL-1438/2020\|EPI_ISL_549220\|2020- | ···················· | ···················· | ····················· | ···················· | ···················· | ···················· |
| J | 3606 | hCoV-19/USA/FL-BPHL-1440/2020\|EPI_ISL_549222\|2020- | ···················· | ···················· | ····················· | ···················· | ···················· | ···················· |
| J | 3607 | hCoV-19/USA/FL-BPHL-1441/2020\|EPI_ISL_549223\|2020- | ···················· | ···················· | ····················· | ···················· | ···················· | ···················· |
| J | 3608 | hCoV-19/USA/FL-BPHL-1442/2020\|EPI_ISL_549224\|2020- | ···················· | ···················· | ····················· | ···················· | ···················· | ···················· |
| J | 3609 | hCoV-19/USA/FL-BPHL-1443/2020\|EPI_ISL_549225\|2020- | ···················· | ·A·················· | ····················· | ···················· | ···················· | ···················· |
| J | 3610 | hCoV-19/USA/FL-BPHL-1444/2020\|EPI_ISL_549226\|2020- | ···················· | ···················· | ····················· | ···················· | ···················· | ···················· |
| J | 3611 | hCoV-19/USA/FL-BPHL-1445/2020\|EPI_ISL_549227\|2020- | ···················· | ···················· | ····················· | ···················· | ···················· | ···················· |
| J | 3612 | hCoV-19/USA/FL-BPHL-1449/2020\|EPI_ISL_549231\|2020- | ···················· | ···················· | ····················· | ···················· | ···················· | ···················· |
| J | 3613 | hCoV-19/USA/FL-BPHL-1450/2020\|EPI_ISL_549232\|2020- | ···················· | ···················· | ····················· | ···················· | ···················· | ···················· |
| J | 3614 | hCoV-19/USA/FL-BPHL-1454/2020\|EPI_ISL_549236\|2020- | ···················· | ···················· | ····················· | ···················· | ···················· | ···················· |
| J | 3615 | hCoV-19/USA/FL-BPHL-1455/2020\|EPI_ISL_549237\|2020- | ···················· | ···················· | ····················· | ···················· | ···················· | ···················· |
| J | 3616 | hCoV-19/USA/FL-BPHL-1456/2020\|EPI_ISL_549238\|2020- | ···················· | ···················· | ····················· | ···················· | ···················· | ···················· |
| J | 3617 | hCoV-19/USA/FL-BPHL-1457/2020\|EPI_ISL_549239\|2020- | ···················· | ···················· | ····················· | ···················· | ···················· | ···················· |
| J | 3618 | hCoV-19/USA/FL-BPHL-1458/2020\|EPI_ISL_549240\|2020- | ···················· | ···················· | ····················· | ···················· | ···················· | ···················· |
| J | 3620 | hCoV-19/USA/FL-BPHL-1463/2020\|EPI_ISL_549245\|2020- | ···················· | ···················· | ····················· | ···················· | ···················· | ···················· |
| J | 3621 | hCoV-19/USA/FL-BPHL-1465/2020\|EPI_ISL_549247\|2020- | ···················· | ···················· | ····················· | ···················· | ···················· | ···················· |
| J | 3622 | hCoV-19/USA/FL-BPHL-1466/2020\|EPI_ISL_549248\|2020- | ···················· | ···················· | ····················· | ···················· | ···················· | ···················· |
| J | 3623 | hCoV-19/USA/FL-BPHL-1467/2020\|EPI_ISL_549249\|2020- | ···················· | ···················· | ····················· | ···················· | ···················· | ···················· |
| J | 3624 | hCoV-19/USA/FL-BPHL-1468/2020\|EPI_ISL_549250\|2020- | ···················· | ···················· | ····················· | ···················· | ···················· | ···················· |
| J | 3625 | hCoV-19/USA/FL-BPHL-1469/2020\|EPI_ISL_549251\|2020- | ···················· | ···················· | ····················· | ···················· | ···················· | ···················· |
| J | 3626 | hCoV-19/USA/FL-BPHL-1471/2020\|EPI_ISL_549253\|2020- | ···················· | ···················· | ····················· | ···················· | ···················· | ···················· |
| J | 3627 | hCoV-19/USA/FL-BPHL-1473/2020\|EPI_ISL_549255\|2020- | ···················· | ···················· | ····················· | ···················· | ···················· | ···················· |
| J | 3628 | hCoV-19/USA/FL-BPHL-1476/2020\|EPI_ISL_549258\|2020- | ···················· | ···················· | ····················· | ···················· | ···················· | ···················· |
| J | 3629 | hCoV-19/USA/FL-BPHL-1477/2020\|EPI_ISL_549259\|2020- | ···················· | ···················· | ····················· | ···················· | ···················· | ···················· |
| J | 3630 | hCoV-19/England/MILK-9A8326/2020\|EPI_ISL_550200\|20 | ···················· | ···················· | ····················· | ···················· | ···················· | ···················· |
| J | 3631 | hCoV-19/England/MILK-9A6AC9/2020\|EPI_ISL_550201\|20 | ···················· | ···················· | ····················· | ···················· | ···················· | ···················· |
| J | 3632 | hCoV-19/England/MILK-9A862D/2020\|EPI_ISL_550203\|20 | ···················· | ···················· | ····················· | ···················· | ···················· | ···················· |
| J | 3633 | hCoV-19/England/MILK-9A5101/2020\|EPI_ISL_550204\|20 | ···················· | ···················· | ····················· | ···················· | ···················· | ···················· |
| J | 3634 | hCoV-19/England/MILK-9A8D28/2020\|EPI_ISL_550205\|20 | ···················· | ···················· | ····················· | ···················· | ···················· | ···················· |
| J | 3635 | hCoV-19/England/MILK-9A8308/2020\|EPI_ISL_550206\|20 | ···················· | ···················· | ····················· | ···················· | ···················· | ···················· |
| J | 3636 | hCoV-19/England/MILK-9A559C/2020\|EPI_ISL_550207\|20 | ···················· | ···················· | ····················· | ···················· | ···················· | ···················· |
| J | 3637 | hCoV-19/England/MILK-9A8CC1/2020\|EPI_ISL_550208\|20 | ···················· | ····A··············· | ····················· | ···················· | ···················· | ···················· |
| J | 3638 | hCoV-19/England/MILK-9A56A8/2020\|EPI_ISL_550211\|20 | ···················· | ···················· | ····················· | ···················· | ···················· | ···················· |
| J | 3639 | hCoV-19/England/MILK-9A8C0D/2020\|EPI_ISL_550212\|20 | ···················· | ···················· | ····················· | ···················· | ···················· | ···················· |
| J | 3640 | hCoV-19/England/MILK-9A6BF3/2020\|EPI_ISL_550213\|20 | ···················· | ···················· | ····················· | ···················· | ···················· | ···················· |
| J | 3641 | hCoV-19/England/MILK-9A8CEF/2020\|EPI_ISL_550214\|20 | ···················· | ···················· | ····················· | ···················· | ···················· | ···················· |
| J | 3642 | hCoV-19/England/MILK-9A8D55/2020\|EPI_ISL_550215\|20 | ···················· | ···················· | ····················· | ···················· | ···················· | ···················· |
| J | 3643 | hCoV-19/England/MILK-9A8C49/2020\|EPI_ISL_550216\|20 | ···················· | ···················· | ····················· | ···················· | ···················· | ···················· |
| J | 3644 | hCoV-19/England/MILK-9A66F2/2020\|EPI_ISL_550217\|20 | ···················· | ···················· | ····················· | ···················· | ···················· | ···················· |
| J | 3645 | hCoV-19/England/MILK-9A84C9/2020\|EPI_ISL_550218\|20 | ···················· | ···················· | ····················· | ···················· | ···················· | ···················· |
| J | 3646 | hCoV-19/England/MILK-9A5769/2020\|EPI_ISL_550219\|20 | ···················· | ···················· | ····················· | ···················· | ···················· | ···················· |
| J | 3647 | hCoV-19/England/MILK-9A6C4B/2020\|EPI_ISL_550220\|20 | ···················· | ···················· | ····················· | ···················· | ···················· | ···················· |
| J | 3648 | hCoV-19/England/MILK-9A6883/2020\|EPI_ISL_550221\|20 | ···················· | ···················· | ····················· | ···················· | ···················· | ···················· |
| J | 3649 | hCoV-19/England/MILK-9A566C/2020\|EPI_ISL_550222\|20 | ···················· | ···················· | ····················· | ···················· | ···················· | ···················· |
| J | 3650 | hCoV-19/England/MILK-9A6874/2020\|EPI_ISL_550223\|20 | ···················· | ···················· | ····················· | ···················· | ···················· | ···················· |
| J | 3651 | hCoV-19/England/MILK-9A51E3/2020\|EPI_ISL_550224\|20 | ···················· | ···················· | ····················· | ···················· | ···················· | ···················· |
| J | 3652 | hCoV-19/England/MILK-9A8DA0/2020\|EPI_ISL_550225\|20 | ···················· | ···················· | ····················· | ···················· | ···················· | ···················· |
| J | 3653 | hCoV-19/England/MILK-9A6D66/2020\|EPI_ISL_550227\|20 | ···················· | ·A·················· | ····················· | ···················· | ···················· | ···A················ |
| J | 3654 | hCoV-19/England/MILK-9A6E27/2020\|EPI_ISL_550228\|20 | ···················· | ···················· | ····················· | ···················· | ···················· | ···················· |
| J | 3655 | hCoV-19/England/MILK-9A68DE/2020\|EPI_ISL_550229\|20 | ···················· | ···················· | ····················· | ···················· | ···················· | ···················· |
| J | 3656 | hCoV-19/England/MILK-9A66C5/2020\|EPI_ISL_550231\|20 | ···················· | ···················· | ····················· | ···················· | ···················· | ···················· |
| J | 3657 | hCoV-19/England/MILK-9A6A50/2020\|EPI_ISL_550232\|20 | ···················· | ···················· | ····················· | ···················· | ···················· | ···················· |
| J | 3658 | hCoV-19/England/MILK-9A8599/2020\|EPI_ISL_550233\|20 | ···················· | ···················· | ····················· | ···················· | ···················· | ···················· |
| J | 3659 | hCoV-19/England/MILK-9A69AE/2020\|EPI_ISL_550234\|20 | ···················· | ···················· | ····················· | ···················· | ···················· | ···················· |
| J | 3660 | hCoV-19/England/MILK-9A5417/2020\|EPI_ISL_550235\|20 | ···················· | ·A·················· | ····················· | ···················· | ···················· | ···A················ |

| | | | | | | | | |
|---|---|---|---|---|---|---|---|---|
| J | 3661 | hCoV-19/England/MILK-9A8317/2020\|EPI_ISL_550236\|20 | .................... | .................... | .................... | .................... | .................... | .................... |
| J | 3662 | hCoV-19/England/MILK-9A54EA/2020\|EPI_ISL_550238\|20 | .................... | .................... | .................... | .................... | .................... | .................... |
| J | 3663 | hCoV-19/England/MILK-9A515C/2020\|EPI_ISL_550239\|20 | .................... | .A.................. | .................... | .................... | .................... | .................... |
| J | 3664 | hCoV-19/England/MILK-9A6DA2/2020\|EPI_ISL_550240\|20 | .................... | .A......A........... | .................... | .................... | .................... | .................... |
| J | 3665 | hCoV-19/England/MILK-9A8AA9/2020\|EPI_ISL_550241\|20 | .................... | .A......A........... | .................... | .................... | .................... | .................... |
| J | 3666 | hCoV-19/England/MILK-9A681A/2020\|EPI_ISL_550243\|20 | .................... | .................... | .................... | .................... | .................... | .................... |
| J | 3667 | hCoV-19/England/MILK-999C78/2020\|EPI_ISL_550266\|20 | .................... | .................... | .................... | .................... | .................... | .................... |
| J | 3668 | hCoV-19/England/MILK-99A13C/2020\|EPI_ISL_550267\|20 | .................... | .................... | .................... | .................... | .................... | .................... |
| J | 3669 | hCoV-19/England/MILK-999E90/2020\|EPI_ISL_550268\|20 | .................... | .................... | .................... | .................... | .................... | .................... |
| J | 3670 | hCoV-19/England/MILK-99A381/2020\|EPI_ISL_550269\|20 | .................... | .................... | .................... | .................... | .................... | .................... |
| J | 3671 | hCoV-19/England/MILK-999FD9/2020\|EPI_ISL_550270\|20 | .................... | .................... | .................... | .................... | .................... | .................... |
| J | 3672 | hCoV-19/England/MILK-99A73A/2020\|EPI_ISL_550272\|20 | .................... | .................... | .................... | .................... | .................... | .................... |
| J | 3673 | hCoV-19/England/MILK-999F24/2020\|EPI_ISL_550276\|20 | .................... | .................... | .................... | .................... | .................... | .................... |
| J | 3674 | hCoV-19/England/MILK-99A4BB/2020\|EPI_ISL_550278\|20 | .................... | .................... | .................... | .................... | .................... | .................... |
| J | 3675 | hCoV-19/England/MILK-999E36/2020\|EPI_ISL_550279\|20 | .................... | .................... | .................... | .................... | .................... | .................... |
| J | 3676 | hCoV-19/England/MILK-999F51/2020\|EPI_ISL_550280\|20 | .................... | .................... | .................... | .................... | .................... | .................... |
| J | 3677 | hCoV-19/England/MILK-99A7A3/2020\|EPI_ISL_550281\|20 | .................... | .................... | .................... | .................... | .................... | .................... |
| J | 3678 | hCoV-19/England/MILK-99A5D6/2020\|EPI_ISL_550282\|20 | .................... | .................... | .................... | .................... | .................... | .................... |
| J | 3679 | hCoV-19/England/MILK-999C2D/2020\|EPI_ISL_550283\|20 | .................... | .................... | .................... | .................... | .................... | .................... |
| J | 3680 | hCoV-19/England/MILK-99A0E4/2020\|EPI_ISL_550285\|20 | .................... | .................... | .................... | .................... | .................... | .................... |
| J | 3681 | hCoV-19/England/MILK-99A099/2020\|EPI_ISL_550286\|20 | .................... | .................... | .................... | .................... | .....T.............. | .................... |
| J | 3682 | hCoV-19/England/MILK-999D66/2020\|EPI_ISL_550287\|20 | .................... | .................... | .................... | .................... | .................... | .................... |
| J | 3683 | hCoV-19/England/MILK-99A8BF/2020\|EPI_ISL_550289\|20 | .................... | .................... | .................... | .................... | .................... | .................... |
| J | 3684 | hCoV-19/England/MILK-999FBB/2020\|EPI_ISL_550290\|20 | .................... | .................... | .................... | .................... | .................... | .................... |
| J | 3685 | hCoV-19/England/MILK-99A600/2020\|EPI_ISL_550291\|20 | .................... | .................... | .................... | .................... | .................... | .................... |
| J | 3686 | hCoV-19/England/MILK-999CA5/2020\|EPI_ISL_550292\|20 | .................... | .................... | .................... | .................... | .................... | .................... |
| J | 3687 | hCoV-19/England/MILK-99A372/2020\|EPI_ISL_550293\|20 | .................... | .................... | .................... | .................... | .................... | .................... |
| J | 3688 | hCoV-19/England/MILK-99A749/2020\|EPI_ISL_550294\|20 | .................... | .................... | .................... | .................... | .................... | .................... |
| J | 3689 | hCoV-19/England/MILK-99A58B/2020\|EPI_ISL_550295\|20 | .................... | .................... | .................... | .................... | .................... | .................... |
| J | 3690 | hCoV-19/England/MILK-999D93/2020\|EPI_ISL_550297\|20 | .................... | .................... | .................... | .................... | .................... | .................... |
| J | 3691 | hCoV-19/England/MILK-999DDF/2020\|EPI_ISL_550298\|20 | .................... | .................... | .................... | .................... | .....T.............. | .................... |
| J | 3692 | hCoV-19/Scotland/MILK-99A309/2020\|EPI_ISL_550299\|2 | .................... | .................... | .................... | .................... | .................... | .................... |
| J | 3693 | hCoV-19/England/ALDP-92B0E8/2020\|EPI_ISL_551501\|20 | .................... | .................... | .................... | .................... | .................... | .................... |
| J | 3694 | hCoV-19/England/ALDP-924BDF/2020\|EPI_ISL_551503\|20 | .................... | .................... | .................... | .................... | .................... | .................... |
| J | 3695 | hCoV-19/England/ALDP-9009B1/2020\|EPI_ISL_551504\|20 | .................... | .................... | .................... | .................... | .................... | .................... |
| J | 3696 | hCoV-19/England/ALDP-92D17A/2020\|EPI_ISL_551505\|20 | .................... | .A.................. | .................... | .................... | .................... | .................... |
| J | 3697 | hCoV-19/England/ALDP-90C6CD/2020\|EPI_ISL_551506\|20 | .................... | .................... | .................... | .................... | .................... | .................... |
| J | 3698 | hCoV-19/England/ALDP-92D14D/2020\|EPI_ISL_551507\|20 | .................... | .................... | .................... | .................... | .................... | .................... |
| J | 3699 | hCoV-19/England/ALDP-92D1F2/2020\|EPI_ISL_551508\|20 | .................... | .................... | .................... | .................... | .................... | .................... |
| J | 3700 | hCoV-19/England/ALDP-90F0EA/2020\|EPI_ISL_551509\|20 | .................... | .................... | .................... | .................... | .................... | .................... |
| J | 3701 | hCoV-19/England/ALDP-9060D4/2020\|EPI_ISL_551510\|20 | .................... | .................... | .................... | .................... | .................... | .................... |
| J | 3702 | hCoV-19/England/ALDP-90A234/2020\|EPI_ISL_551512\|20 | .................... | .................... | .................... | .................... | .................... | .................... |
| J | 3703 | hCoV-19/England/ALDP-916C46/2020\|EPI_ISL_551514\|20 | .................... | .A.................. | .................... | .................... | .................... | .................... |
| J | 3704 | hCoV-19/England/ALDP-90456E/2020\|EPI_ISL_551515\|20 | .................... | .................... | .................... | .................... | .................... | .................... |
| J | 3705 | hCoV-19/England/ALDP-904B4E/2020\|EPI_ISL_551516\|20 | .................... | .................... | .................... | .................... | .................... | .................... |
| J | 3706 | hCoV-19/England/ALDP-924A2D/2020\|EPI_ISL_551518\|20 | .................... | .................... | .................... | .................... | .................... | .................... |
| J | 3707 | hCoV-19/England/ALDP-90069C/2020\|EPI_ISL_551523\|20 | .................... | .................... | .................... | .................... | .................... | .................... |
| J | 3708 | hCoV-19/England/ALDP-933D40/2020\|EPI_ISL_551525\|20 | .................... | .................... | .................... | .................... | .................... | .................... |
| J | 3709 | hCoV-19/England/ALDP-8FD38D/2020\|EPI_ISL_551526\|20 | .................... | .................... | .................... | .................... | .................... | .................... |
| J | 3710 | hCoV-19/England/ALDP-8FC272/2020\|EPI_ISL_551528\|20 | .................... | .................... | .................... | .................... | .................... | .................... |
| J | 3711 | hCoV-19/England/ALDP-91DA72/2020\|EPI_ISL_551529\|20 | .................... | .................... | .................... | .................... | .................... | .................... |
| J | 3712 | hCoV-19/England/ALDP-9002A7/2020\|EPI_ISL_551531\|20 | .................... | .................... | .................... | .................... | .................... | .................... |
| J | 3713 | hCoV-19/England/ALDP-92E3DD/2020\|EPI_ISL_551532\|20 | .................... | .................... | .................... | .................... | .................... | .................... |
| J | 3714 | hCoV-19/England/ALDP-90C7AC/2020\|EPI_ISL_551533\|20 | .................... | .A.................. | .................... | .................... | .................... | .................... |
| J | 3715 | hCoV-19/England/ALDP-90CDD7/2020\|EPI_ISL_551535\|20 | .................... | .................... | .................... | .................... | .................... | .................... |
| J | 3716 | hCoV-19/England/ALDP-92EA50/2020\|EPI_ISL_551536\|20 | .................... | .A.................. | .................... | .................... | .................... | .................... |
| J | 3717 | hCoV-19/England/ALDP-9060F2/2020\|EPI_ISL_551537\|20 | .................... | .................... | .................... | .................... | .................... | .................... |
| J | 3718 | hCoV-19/England/ALDP-938789/2020\|EPI_ISL_551538\|20 | .................... | .................... | .................... | .................... | .................... | .................... |
| J | 3719 | hCoV-19/England/ALDP-9058A0/2020\|EPI_ISL_551539\|20 | .................... | .................... | .................... | .................... | .................... | .................... |
| J | 3720 | hCoV-19/England/ALDP-924A78/2020\|EPI_ISL_551540\|20 | .................... | .................... | .................... | .................... | .................... | .................... |
| J | 3721 | hCoV-19/England/ALDP-9055C7/2020\|EPI_ISL_551541\|20 | .................... | .................... | .................... | .................... | .................... | .................... |
| J | 3722 | hCoV-19/England/ALDP-924A1E/2020\|EPI_ISL_551542\|20 | .................... | .................... | .................... | .................... | .................... | .................... |
| J | 3723 | hCoV-19/England/ALDP-90E573/2020\|EPI_ISL_551543\|20 | .................... | .................... | .................... | .................... | .................... | .................... |
| J | 3724 | hCoV-19/England/ALDP-925DAB/2020\|EPI_ISL_551544\|20 | .................... | .................... | .................... | .................... | .................... | .................... |
| J | 3725 | hCoV-19/England/ALDP-9342AA/2020\|EPI_ISL_551545\|20 | .................... | .A.................. | .................... | .................... | .................... | .................... |
| J | 3726 | hCoV-19/England/ALDP-908658/2020\|EPI_ISL_551546\|20 | .................... | .................... | .................... | .................... | .................... | .................... |
| J | 3727 | hCoV-19/England/ALDP-90606B/2020\|EPI_ISL_551547\|20 | .................... | .................... | .................... | .................... | .................... | .................... |
| J | 3728 | hCoV-19/England/ALDP-92A0CB/2020\|EPI_ISL_551548\|20 | .................... | .................... | .................... | .................... | .................... | .................... |
| J | 3729 | hCoV-19/England/ALDP-9025BB/2020\|EPI_ISL_551549\|20 | .................... | .................... | .................... | .................... | .................... | .................... |
| J | 3730 | hCoV-19/England/ALDP-8FD8D9/2020\|EPI_ISL_551550\|20 | .................... | .................... | .................... | .................... | .................... | .................... |
| J | 3731 | hCoV-19/England/ALDP-90F3B4/2020\|EPI_ISL_551551\|20 | .................... | .................... | .................... | .................... | .................... | .................... |
| J | 3732 | hCoV-19/England/ALDP-936FFC/2020\|EPI_ISL_551552\|20 | .................... | .................... | .................... | .................... | .................... | .................... |
| J | 3733 | hCoV-19/England/ALDP-924CBE/2020\|EPI_ISL_551553\|20 | .................... | .................... | .................... | .................... | .................... | .................... |
| J | 3734 | hCoV-19/England/ALDP-9050C6/2020\|EPI_ISL_551554\|20 | .................... | .................... | .................... | .................... | .................... | .................... |
| J | 3735 | hCoV-19/England/ALDP-900C4F/2020\|EPI_ISL_551555\|20 | .................... | .................... | .................... | .................... | .................... | .................... |
| J | 3736 | hCoV-19/England/ALDP-91A18F/2020\|EPI_ISL_551556\|20 | .................... | .................... | .................... | .................... | .................... | .................... |
| J | 3737 | hCoV-19/England/ALDP-9087DD/2020\|EPI_ISL_551557\|20 | .................... | .................... | .................... | .................... | .................... | .................... |
| J | 3738 | hCoV-19/England/ALDP-92DF34/2020\|EPI_ISL_551558\|20 | .................... | .................... | .................... | .................... | .................... | .................... |
| J | 3739 | hCoV-19/England/ALDP-92E8FC/2020\|EPI_ISL_551559\|20 | .................... | .A.................. | .................... | .................... | .................... | .................... |
| J | 3740 | hCoV-19/England/ALDP-907765/2020\|EPI_ISL_551560\|20 | .................... | .................... | .................... | .................... | .................... | .................... |

| | | | | | | | | |
|---|---|---|---|---|---|---|---|---|
| J | 3741 | hCoV-19/England/ALDP-90080F/2020|EPI_ISL_551562|20 | ···················· | ···················· | ···················· | ···················· | ···················· | ···················· |
| J | 3742 | hCoV-19/England/ALDP-92400D/2020|EPI_ISL_551563|20 | ···················· | ···················· | ···················· | ···················· | ···················· | ···················· |
| J | 3743 | hCoV-19/England/ALDP-909AF1/2020|EPI_ISL_551564|20 | ···················· | ···················· | ···················· | ···················· | ···················· | ···················· |
| J | 3744 | hCoV-19/England/ALDP-925FD2/2020|EPI_ISL_551565|20 | ···················· | ···················· | ···················· | ···················· | ···················· | ···················· |
| J | 3745 | hCoV-19/England/ALDP-9018C2/2020|EPI_ISL_551567|20 | ···················· | ···················· | ···················· | ···················· | ···················· | ···················· |
| J | 3746 | hCoV-19/England/ALDP-901965/2020|EPI_ISL_551568|20 | ···················· | ···················· | ···················· | ···················· | ···················· | ···················· |
| J | 3747 | hCoV-19/England/ALDP-90671B/2020|EPI_ISL_551569|20 | ···················· | ···················· | ···················· | ···················· | ···················· | ···················· |
| J | 3748 | hCoV-19/England/ALDP-923D16/2020|EPI_ISL_551570|20 | ···················· | ···················· | ···················· | ···················· | ···················· | ···················· |
| J | 3749 | hCoV-19/England/ALDP-90222D/2020|EPI_ISL_551571|20 | ···················· | ···················· | ···················· | ···················· | ···················· | ···················· |
| J | 3750 | hCoV-19/England/ALDP-90A164/2020|EPI_ISL_551572|20 | ···················· | ···················· | ···················· | ···················· | ···················· | ···················· |
| J | 3751 | hCoV-19/England/ALDP-905DCF/2020|EPI_ISL_551573|20 | ···················· | ···················· | ···················· | ···················· | ···················· | ···················· |
| J | 3752 | hCoV-19/England/MILK-9957A8/2020|EPI_ISL_551598|20 | ···················· | ···················· | ···················· | ···················· | ···················· | ···················· |
| J | 3753 | hCoV-19/England/ALDP-928E87/2020|EPI_ISL_552821|20 | ···················· | ···················· | ···················· | ···················· | ···················· | ···················· |
| J | 3754 | hCoV-19/England/ALDP-92D532/2020|EPI_ISL_552822|20 | ···················· | ···················· | ···················· | ···················· | ···················· | ···················· |
| J | 3755 | hCoV-19/England/MILK-93873E/2020|EPI_ISL_552823|20 | ···················· | ···················· | ···················· | ···················· | ···················· | ···················· |
| J | 3756 | hCoV-19/England/ALDP-93719B/2020|EPI_ISL_552824|20 | ···················· | ···················· | ···················· | ···················· | ···················· | ···················· |
| J | 3757 | hCoV-19/England/ALDP-936B07/2020|EPI_ISL_552825|20 | ···················· | ···················· | ···················· | ···················· | ···················· | ···················· |
| J | 3758 | hCoV-19/England/ALDP-920E16/2020|EPI_ISL_552827|20 | ···················· | ·A·················· | ···················· | ···················· | ···················· | ···················· |
| J | 3759 | hCoV-19/England/ALDP-925567/2020|EPI_ISL_552828|20 | ···················· | ···················· | ···················· | ···················· | ···················· | ···················· |
| J | 3760 | hCoV-19/England/ALDP-935B08/2020|EPI_ISL_552829|20 | ···················· | ···················· | ···················· | ···················· | ···················· | ···················· |
| J | 3761 | hCoV-19/England/ALDP-9205A8/2020|EPI_ISL_552830|20 | ···················· | ···················· | ···················· | ···················· | ···················· | ···················· |
| J | 3762 | hCoV-19/England/ALDP-920A30/2020|EPI_ISL_552831|20 | ···················· | ···················· | ···················· | ···················· | ···················· | ···················· |
| J | 3763 | hCoV-19/England/ALDP-90A8AB/2020|EPI_ISL_552832|20 | ···················· | ···················· | ···················· | ···················· | ···················· | ···················· |
| J | 3764 | hCoV-19/England/ALDP-934A48/2020|EPI_ISL_552833|20 | ···················· | ···················· | ···················· | ···················· | ···················· | ···················· |
| J | 3765 | hCoV-19/England/ALDP-92C2B4/2020|EPI_ISL_552834|20 | ···················· | ···················· | ···················· | ···················· | ···················· | ···················· |
| J | 3766 | hCoV-19/England/ALDP-935959/2020|EPI_ISL_552835|20 | ···················· | ···················· | ···················· | ···················· | ···················· | ···················· |
| J | 3767 | hCoV-19/England/ALDP-9031E3/2020|EPI_ISL_552836|20 | ···················· | ···················· | ···················· | ···················· | ···················· | ···················· |
| J | 3768 | hCoV-19/England/ALDP-9355A0/2020|EPI_ISL_552837|20 | ···················· | ···················· | ···················· | ···················· | ···················· | ···················· |
| J | 3769 | hCoV-19/England/ALDP-91D650/2020|EPI_ISL_552839|20 | ···················· | ···················· | ···················· | ···················· | ···················· | ···················· |
| J | 3770 | hCoV-19/England/ALDP-92065A/2020|EPI_ISL_552840|20 | ···················· | ···················· | ···················· | ···················· | ···················· | ···················· |
| J | 3771 | hCoV-19/England/MILK-93F5C4/2020|EPI_ISL_552842|20 | ···················· | ···················· | ···················· | ···················· | ···················· | ···················· |
| J | 3772 | hCoV-19/England/ALDP-925813/2020|EPI_ISL_552843|20 | ···················· | ···················· | ···················· | ···················· | ···················· | ···················· |
| J | 3773 | hCoV-19/England/ALDP-925840/2020|EPI_ISL_552844|20 | ···················· | ···················· | ···················· | ···················· | ···················· | ···················· |
| J | 3774 | hCoV-19/England/ALDP-925725/2020|EPI_ISL_552845|20 | ···················· | ···················· | ···················· | ···················· | ···················· | ···················· |
| J | 3775 | hCoV-19/England/ALDP-93732C/2020|EPI_ISL_552846|20 | ···················· | ···················· | ···················· | ···················· | ···················· | ···················· |
| J | 3776 | hCoV-19/England/ALDP-926AA3/2020|EPI_ISL_552847|20 | ···················· | ···················· | ···················· | ···················· | ···················· | ···················· |
| J | 3777 | hCoV-19/England/ALDP-9257F8/2020|EPI_ISL_552848|20 | ···················· | ···················· | ···················· | ···················· | ···················· | ···················· |
| J | 3778 | hCoV-19/England/ALDP-929CC8/2020|EPI_ISL_552849|20 | ···················· | ···················· | ···················· | ···················· | ···················· | ···················· |
| J | 3779 | hCoV-19/England/ALDP-918EE4/2020|EPI_ISL_552850|20 | ···················· | ···················· | ···················· | ···················· | ···················· | ···················· |
| J | 3780 | hCoV-19/England/ALDP-925761/2020|EPI_ISL_552851|20 | ···················· | ···················· | ···················· | ···················· | ···················· | ···················· |
| J | 3781 | hCoV-19/England/MILK-93D13B/2020|EPI_ISL_552852|20 | ···················· | ···················· | ···················· | ···················· | ···················· | ···················· |
| J | 3782 | hCoV-19/England/ALDP-920B1F/2020|EPI_ISL_552853|20 | ···················· | ···················· | ···················· | ···················· | ···················· | ···················· |
| J | 3783 | hCoV-19/England/ALDP-902A70/2020|EPI_ISL_552855|20 | ···················· | ···················· | ···················· | ···················· | ···················· | ···················· |
| J | 3784 | hCoV-19/England/ALDP-926C07/2020|EPI_ISL_552856|20 | ···················· | ···················· | ···················· | ···················· | ···················· | ···················· |
| J | 3785 | hCoV-19/England/ALDP-920B2E/2020|EPI_ISL_552857|20 | ···················· | ···················· | ···················· | ···················· | ···················· | ···················· |
| J | 3786 | hCoV-19/England/ALDP-90321D/2020|EPI_ISL_552858|20 | ···················· | ···················· | ···················· | ···················· | ···················· | ···················· |
| J | 3787 | hCoV-19/England/ALDP-9358E3/2020|EPI_ISL_552860|20 | ···················· | ···················· | ···················· | ···················· | ···················· | ···················· |
| J | 3788 | hCoV-19/England/ALDP-923E9B/2020|EPI_ISL_552861|20 | ···················· | ···················· | ···················· | ···················· | ···················· | ···················· |
| J | 3789 | hCoV-19/England/ALDP-9255A3/2020|EPI_ISL_552862|20 | ···················· | ···················· | ···················· | ···················· | ···················· | ···················· |
| J | 3790 | hCoV-19/England/ALDP-92099D/2020|EPI_ISL_552863|20 | ···················· | ·A·················· | ···················· | ···················· | ···················· | ···················· |
| J | 3791 | hCoV-19/England/ALDP-92CAF8/2020|EPI_ISL_552864|20 | ···················· | ···················· | ···················· | ···················· | ···················· | ···················· |
| J | 3792 | hCoV-19/England/ALDP-935625/2020|EPI_ISL_552865|20 | ···················· | ···················· | ···················· | ···················· | ···················· | ···················· |
| J | 3793 | hCoV-19/England/ALDP-928AED/2020|EPI_ISL_552866|20 | ···················· | ···················· | ···················· | ···················· | ···················· | ···················· |
| J | 3794 | hCoV-19/England/MILK-93D11D/2020|EPI_ISL_552867|20 | ···················· | ···················· | ···················· | ···················· | ···················· | ···················· |
| J | 3795 | hCoV-19/England/ALDP-920A12/2020|EPI_ISL_552868|20 | ···················· | ·A·················· | ···················· | ···················· | ···················· | ···················· |
| J | 3796 | hCoV-19/England/ALDP-92926C/2020|EPI_ISL_552870|20 | ···················· | ···················· | ···················· | ···················· | ···················· | ···················· |
| J | 3797 | hCoV-19/England/ALDP-929FB0/2020|EPI_ISL_552871|20 | ···················· | ···················· | ···················· | ···················· | ···················· | ···················· |
| J | 3798 | hCoV-19/England/ALDP-903875/2020|EPI_ISL_552872|20 | ···················· | ···················· | ···················· | ···················· | ···················· | ···················· |
| J | 3799 | hCoV-19/England/ALDP-920C85/2020|EPI_ISL_552873|20 | ···················· | ·A·················· | ···················· | ···················· | ···················· | ···················· |
| J | 3800 | hCoV-19/England/MILK-93D502/2020|EPI_ISL_552874|20 | ···················· | ···················· | ···················· | ···················· | ···················· | ···················· |
| J | 3801 | hCoV-19/England/MILK-93DE34/2020|EPI_ISL_552875|20 | ···················· | ···················· | ···················· | ···················· | ···················· | ···················· |
| J | 3802 | hCoV-19/England/ALDP-920960/2020|EPI_ISL_552878|20 | ···················· | ·A·················· | ···················· | ···················· | ···················· | ···················· |
| J | 3803 | hCoV-19/England/MILK-93BA32/2020|EPI_ISL_552879|20 | ···················· | ···················· | ···················· | ···················· | ···················· | ···················· |
| J | 3804 | hCoV-19/England/ALDP-937C7C/2020|EPI_ISL_552881|20 | ···················· | ···················· | ···················· | ···················· | ···················· | ···················· |
| J | 3805 | hCoV-19/England/ALDP-92AACD/2020|EPI_ISL_552882|20 | ···················· | ···················· | ···················· | ···················· | ···················· | ···················· |
| J | 3806 | hCoV-19/England/ALDP-920AE5/2020|EPI_ISL_552883|20 | ···················· | ·A·················· | ···················· | ···················· | ···················· | ···················· |
| J | 3807 | hCoV-19/England/ALDP-91FF44/2020|EPI_ISL_552884|20 | ···················· | ···················· | ···················· | ···················· | ···················· | ···················· |
| J | 3808 | hCoV-19/England/ALDP-928DC6/2020|EPI_ISL_552885|20 | ···················· | ···················· | ···················· | ···················· | ···················· | ···················· |
| J | 3809 | hCoV-19/England/ALDP-930217/2020|EPI_ISL_552886|20 | ···················· | ···················· | ···················· | ···················· | ···················· | ···················· |
| J | 3810 | hCoV-19/England/ALDP-91D22F/2020|EPI_ISL_552887|20 | ···················· | ···················· | ···················· | ···················· | ···················· | ···················· |
| J | 3811 | hCoV-19/England/ALDP-92C560/2020|EPI_ISL_552888|20 | ···················· | ···················· | ···················· | ···················· | ···················· | ···················· |
| J | 3812 | hCoV-19/England/ALDP-903286/2020|EPI_ISL_552889|20 | ···················· | ···················· | ···················· | ···················· | ···················· | ···················· |
| J | 3813 | hCoV-19/England/ALDP-929DF2/2020|EPI_ISL_552890|20 | ···················· | ···················· | ···················· | ···················· | ···················· | ···················· |
| J | 3814 | hCoV-19/England/ALDP-9214C8/2020|EPI_ISL_552891|20 | ···················· | ···················· | ···················· | ···················· | ···················· | ···················· |
| J | 3815 | hCoV-19/England/ALDP-92138F/2020|EPI_ISL_552892|20 | ···················· | ···················· | ···················· | ···················· | ···················· | ···················· |
| J | 3816 | hCoV-19/England/ALDP-920AF4/2020|EPI_ISL_552893|20 | ···················· | ·A·················· | ···················· | ···················· | ···················· | ···················· |
| J | 3817 | hCoV-19/England/ALDP-91D68D/2020|EPI_ISL_552894|20 | ···················· | ···················· | ···················· | ···················· | ···················· | ···················· |
| J | 3818 | hCoV-19/England/ALDP-907D09/2020|EPI_ISL_552895|20 | ···················· | ···················· | ···················· | ···················· | ···················· | ···················· |
| J | 3819 | hCoV-19/England/ALDP-937016/2020|EPI_ISL_552896|20 | ···················· | ···················· | ···················· | ···················· | ···················· | ···················· |
| J | 3820 | hCoV-19/England/ALDP-935BBD/2020|EPI_ISL_552899|20 | ···················· | ···················· | ···················· | ···················· | ···················· | ···················· |

| | | | | | | | |
|---|---|---|---|---|---|---|---|
| J | 3821 hCoV-19/Northern Ireland/MILK-95D9A6/2020\|EPI_ISL_ | ···················· | ······A············· | ···················· | ···················· | ···················· | ···················· |
| J | 3822 hCoV-19/England/ALDP-94ECAF/2020\|EPI_ISL_556234\|20 | ···················· | ···················· | ···················· | ···················· | ···················· | ···················· |
| J | 3823 hCoV-19/Wales/MILK-95DA49/2020\|EPI_ISL_556235\|2020 | ···················· | ···················· | ···················· | ···················· | ···················· | ···················· |
| J | 3824 hCoV-19/England/MILK-6425A3/2020\|EPI_ISL_556237\|20 | ···················· | ···················· | ···················· | ···················· | ···················· | ···················· |
| J | 3825 hCoV-19/England/ALDP-6C565A/2020\|EPI_ISL_556238\|20 | ···················· | ···················· | ···················· | ···················· | ···················· | ···················· |
| J | 3826 hCoV-19/England/ALDP-70B086/2020\|EPI_ISL_557563\|20 | ···················· | ···················· | ···················· | ···················· | ···················· | ···················· |
| J | 3827 hCoV-19/England/ALDP-70409C/2020\|EPI_ISL_557564\|20 | ···················· | ···················· | ···················· | ···················· | ···················· | ···················· |
| J | 3828 hCoV-19/England/ALDP-704EFC/2020\|EPI_ISL_557565\|20 | ···················· | ···················· | ···················· | ···················· | ···················· | ···················· |
| J | 3829 hCoV-19/England/ALDP-6E9052/2020\|EPI_ISL_557566\|20 | ···················· | ···················· | ···················· | ···················· | ···················· | ···················· |
| J | 3830 hCoV-19/England/ALDP-6E261B/2020\|EPI_ISL_557567\|20 | ···················· | ···················· | ···················· | ···················· | ···················· | ···················· |
| J | 3831 hCoV-19/England/ALDP-6EA6C8/2020\|EPI_ISL_557568\|20 | ···················· | ···················· | ···················· | ···················· | ···················· | ···················· |
| J | 3832 hCoV-19/England/ALDP-7004EF/2020\|EPI_ISL_557569\|20 | ···················· | ···················· | ···················· | ···················· | ···················· | ···················· |
| J | 3833 hCoV-19/England/ALDP-703437/2020\|EPI_ISL_557702\|20 | ···················· | ···················· | ···················· | ···················· | ···················· | ···················· |
| J | 3834 hCoV-19/England/ALDP-6EAB14/2020\|EPI_ISL_557703\|20 | ···················· | ···················· | ···················· | ···················· | ···················· | ···················· |
| J | 3835 hCoV-19/England/ALDP-71ED6B/2020\|EPI_ISL_557705\|20 | ···················· | ···················· | ···················· | ···················· | ···················· | ···················· |
| J | 3836 hCoV-19/England/ALDP-70956B/2020\|EPI_ISL_557706\|20 | ···················· | ···················· | ···················· | ···················· | ···················· | ···················· |
| J | 3837 hCoV-19/England/ALDP-7211B4/2020\|EPI_ISL_557707\|20 | ···················· | ···················· | ···················· | ···················· | ···················· | ···················· |
| J | 3838 hCoV-19/England/ALDP-70A01E/2020\|EPI_ISL_557708\|20 | ···················· | ···················· | ···················· | ···················· | ···················· | ···················· |
| J | 3839 hCoV-19/England/ALDP-704DB3/2020\|EPI_ISL_557710\|20 | ···················· | ···················· | ···················· | ···················· | ···················· | ···················· |
| J | 3840 hCoV-19/England/ALDP-708951/2020\|EPI_ISL_557712\|20 | ···················· | ···················· | ···················· | ···················· | ···················· | ···················· |
| J | 3841 hCoV-19/Norway/3117/2020\|EPI_ISL_549140\|2020-08-06 | ···················· | ···················· | ···················· | ···················· | ···················· | ···················· |
| J | 3842 hCoV-19/Norway/3118/2020\|EPI_ISL_549141\|2020-08-06 | ···················· | ···················· | ···················· | ···················· | ···················· | ···················· |
| J | 3843 hCoV-19/Norway/3119/2020\|EPI_ISL_549142\|2020-08-06 | ···················· | ···················· | ···················· | ···················· | ···················· | ···················· |
| J | 3844 hCoV-19/Norway/2949/2020\|EPI_ISL_549143\|2020-07-31 | ···················· | ···················· | ···················· | ···················· | ···················· | ···················· |
| J | 3845 hCoV-19/Norway/3120/2020\|EPI_ISL_549144\|2020-08-06 | ···················· | ···················· | ···················· | ···················· | ···················· | ···················· |
| J | 3846 hCoV-19/Norway/3121/2020\|EPI_ISL_549145\|2020-08-06 | ···················· | ···················· | ···················· | ···················· | ···················· | ···················· |
| J | 3847 hCoV-19/Norway/3123/2020\|EPI_ISL_549146\|2020-08-07 | ···················· | ···················· | ···················· | ···················· | ···················· | ···················· |
| J | 3848 hCoV-19/Norway/3124/2020\|EPI_ISL_549147\|2020-08-07 | ···················· | ···················· | ···················· | ···················· | ···················· | ···················· |
| J | 3849 hCoV-19/Norway/3125/2020\|EPI_ISL_549148\|2020-08-07 | ···················· | ···················· | ···················· | ···················· | ···················· | ···················· |
| J | 3850 hCoV-19/Norway/3126/2020\|EPI_ISL_549149\|2020-08-07 | ···················· | ···················· | ···················· | ···················· | ···················· | ···················· |
| J | 3851 hCoV-19/England/MILK-9A670E/2020\|EPI_ISL_550090\|20 | ···················· | ···················· | ···················· | ···················· | ···················· | ···················· |
| J | 3852 hCoV-19/England/MILK-9A6E81/2020\|EPI_ISL_550091\|20 | ···················· | ···················· | ···················· | ···················· | ···················· | ···················· |
| J | 3853 hCoV-19/England/MILK-9A673B/2020\|EPI_ISL_550092\|20 | ···················· | ···················· | ···················· | ···················· | ···················· | ···················· |
| J | 3854 hCoV-19/England/MILK-9A6EAF/2020\|EPI_ISL_550093\|20 | ···················· | ···················· | ···················· | ···················· | ······T············· | ···················· |
| J | 3855 hCoV-19/England/MILK-9A55F6/2020\|EPI_ISL_550094\|20 | ···················· | ···················· | ···················· | ···················· | ···················· | ···················· |
| J | 3856 hCoV-19/England/MILK-9A5602/2020\|EPI_ISL_550095\|20 | ···················· | ···················· | ···················· | ···················· | ···················· | ···················· |
| J | 3857 hCoV-19/England/MILK-9A52B3/2020\|EPI_ISL_550097\|20 | ···················· | ·A·················· | ···················· | ···················· | ···················· | ···A················ |
| J | 3858 hCoV-19/England/MILK-9A6C2D/2020\|EPI_ISL_550098\|20 | ···················· | ···················· | ···················· | ···················· | ···················· | ···················· |
| J | 3859 hCoV-19/England/MILK-9A6D93/2020\|EPI_ISL_550099\|20 | ···················· | ···················· | ···················· | ···················· | ···················· | ···················· |
| J | 3860 hCoV-19/England/MILK-9A8AE5/2020\|EPI_ISL_550123\|20 | ···················· | ·A·······A·········· | ···················· | ···················· | ···················· | ···················· |
| J | 3861 hCoV-19/England/MILK-9A6DB1/2020\|EPI_ISL_550129\|20 | ···················· | ···················· | ···················· | ···················· | ···················· | ···················· |
| J | 3862 hCoV-19/England/MILK-9A6EFA/2020\|EPI_ISL_550130\|20 | ···················· | ···················· | ···················· | ···················· | ···················· | ···················· |
| J | 3863 hCoV-19/England/MILK-9A853F/2020\|EPI_ISL_550131\|20 | ···················· | ···················· | ···················· | ···················· | ···················· | ···················· |
| J | 3864 hCoV-19/England/MILK-9A8432/2020\|EPI_ISL_550133\|20 | ···················· | ···················· | ···················· | ···················· | ···················· | ···················· |
| J | 3865 hCoV-19/England/ALDP-92BBBA/2020\|EPI_ISL_551455\|20 | ···················· | ···················· | ···················· | ···················· | ···················· | ···················· |
| J | 3866 hCoV-19/England/ALDP-917A78/2020\|EPI_ISL_551458\|20 | ···················· | ···················· | ···················· | ···················· | ···················· | ···················· |
| J | 3867 hCoV-19/England/ALDP-91AF94/2020\|EPI_ISL_551459\|20 | ···················· | ···················· | ···················· | ···················· | ···················· | ···················· |
| J | 3868 hCoV-19/England/ALDP-92AF37/2020\|EPI_ISL_551460\|20 | ···················· | ·A·················· | ···················· | ···················· | ···················· | ···················· |
| J | 3869 hCoV-19/England/ALDP-90F624/2020\|EPI_ISL_551461\|20 | ···················· | ···················· | ···················· | ···················· | ···················· | ···················· |
| J | 3870 hCoV-19/England/ALDP-904BD5/2020\|EPI_ISL_551462\|20 | ···················· | ···················· | ···················· | ···················· | ···················· | ···················· |
| J | 3871 hCoV-19/England/ALDP-90C79D/2020\|EPI_ISL_551463\|20 | ···················· | ···················· | ···················· | ···················· | ···················· | ···················· |
| J | 3872 hCoV-19/England/ALDP-904795/2020\|EPI_ISL_551464\|20 | ···················· | ···················· | ···················· | ···················· | ···················· | ···················· |
| J | 3873 hCoV-19/England/MILK-975F6A/2020\|EPI_ISL_552789\|20 | ···················· | ·A·················· | ···················· | ···················· | ···················· | ···A················ |
| J | 3874 hCoV-19/England/MILK-976770/2020\|EPI_ISL_552790\|20 | ···················· | ···················· | ···················· | ···················· | ···················· | ···················· |
| J | 3875 hCoV-19/England/MILK-975CDC/2020\|EPI_ISL_552791\|20 | ···················· | ···················· | ···················· | ···················· | ···················· | ···················· |
| J | 3876 hCoV-19/England/ALDP-981D70/2020\|EPI_ISL_552794\|20 | ···················· | ···················· | ···················· | ···················· | ···················· | ···················· |
| J | 3877 hCoV-19/England/MILK-9768F5/2020\|EPI_ISL_552795\|20 | ···················· | ···················· | ···················· | ···················· | ···················· | ···················· |
| J | 3878 hCoV-19/England/ALDP-6CE24E/2020\|EPI_ISL_556200\|20 | ···················· | ···················· | ···················· | ···················· | ···················· | ···················· |
| J | 3879 hCoV-19/England/ALDP-6CC486/2020\|EPI_ISL_556202\|20 | ···················· | ···················· | ···················· | ···················· | ···················· | ···················· |
| J | 3880 hCoV-19/England/ALDP-94EF1F/2020\|EPI_ISL_556203\|20 | ···················· | ···················· | ···················· | ···················· | ···················· | ···················· |
| J | 3881 hCoV-19/England/MILK-63E7C2/2020\|EPI_ISL_556204\|20 | ···················· | ···················· | ···················· | ···················· | ···················· | ···················· |
| J | 3882 hCoV-19/England/ALDP-639AEC/2020\|EPI_ISL_556206\|20 | ···················· | ···················· | ···················· | ···················· | ···················· | ···················· |
| J | 3883 hCoV-19/England/MILK-95D830/2020\|EPI_ISL_556208\|20 | ···················· | ···················· | ···················· | ···················· | ···················· | ···················· |
| J | 3884 hCoV-19/England/ALDP-6C8C37/2020\|EPI_ISL_556209\|20 | ···················· | ···················· | ···················· | ···················· | ···················· | ···················· |
| J | 3885 hCoV-19/England/ALDP-6C4C2C/2020\|EPI_ISL_556210\|20 | ···················· | ···················· | ···················· | ···················· | ···················· | ···················· |
| J | 3886 hCoV-19/England/MILK-95D900/2020\|EPI_ISL_556211\|20 | ···················· | ·A·················· | ···················· | ···················· | ···················· | ···················· |
| J | 3887 hCoV-19/England/ALDP-6CD397/2020\|EPI_ISL_556212\|20 | ···················· | ···················· | ···················· | ···················· | ···················· | ···················· |
| J | 3888 hCoV-19/England/MILK-95D988/2020\|EPI_ISL_556214\|20 | ···················· | ·············A······ | ···················· | ···················· | ···················· | ···················· |
| J | 3889 hCoV-19/England/ALDP-6CE92B/2020\|EPI_ISL_556215\|20 | ···················· | ···················· | ···················· | ···················· | ···················· | ···················· |
| J | 3890 hCoV-19/England/ALDP-6D3176/2020\|EPI_ISL_556216\|20 | ···················· | ···················· | ···················· | ···················· | ···················· | ···················· |
| J | 3891 hCoV-19/England/ALDP-6D67DB/2020\|EPI_ISL_556217\|20 | ···················· | ···················· | ···················· | ···················· | ···················· | ···················· |
| J | 3892 hCoV-19/England/ALDP-6D34D7/2020\|EPI_ISL_556219\|20 | ···················· | ···················· | ···················· | ···················· | ···················· | ···················· |
| J | 3893 hCoV-19/England/ALDP-6C753D/2020\|EPI_ISL_556220\|20 | ···················· | ···················· | ···················· | ···················· | ···················· | ···················· |
| J | 3894 hCoV-19/England/ALDP-63D15C/2020\|EPI_ISL_556221\|20 | ···················· | ···················· | ···················· | ···················· | ···················· | ···················· |
| J | 3895 hCoV-19/England/ALDP-6D4351/2020\|EPI_ISL_556222\|20 | ···················· | ···················· | ···················· | ···················· | ···················· | ···················· |
| J | 3896 hCoV-19/England/MILK-6426BF/2020\|EPI_ISL_556223\|20 | ···················· | ···················· | ···················· | ···················· | ···················· | ···················· |
| J | 3897 hCoV-19/England/ALDP-6CB6BE/2020\|EPI_ISL_556225\|20 | ···················· | ···················· | ···················· | ···················· | ···················· | ···················· |
| J | 3898 hCoV-19/Wales/ALDP-6C3E72/2020\|EPI_ISL_556227\|2020 | ···················· | ···················· | ···················· | ···················· | ···················· | ···················· |
| J | 3899 hCoV-19/England/ALDP-6D29AC/2020\|EPI_ISL_556228\|20 | ···················· | ···················· | ···················· | ···················· | ···················· | ···················· |
| J | 3900 hCoV-19/England/ALDP-6C60B5/2020\|EPI_ISL_556229\|20 | ···················· | ···················· | ···················· | ···················· | ···················· | ···················· |

| | | | | | | | | |
|---|---|---|---|---|---|---|---|---|
| J | 3901 | hCoV-19/England/ALDP-6D68D8/2020\|EPI_ISL_556230\|20 | ···················· | ···················· | ···················· | ···················· | ···················· | ···················· |
| J | 3902 | hCoV-19/England/ALDP-94EAF0/2020\|EPI_ISL_556231\|20 | ···················· | ···················· | ···················· | ···················· | ···················· | ···················· |
| J | 3903 | hCoV-19/England/MILK-95D8F4/2020\|EPI_ISL_556232\|20 | ···················· | ·A·················· | ···················· | ···················· | ·Y·················· | ···················· |
| J | 3904 | hCoV-19/England/ALDP-6D86CD/2020\|EPI_ISL_556239\|20 | ···················· | ···················· | ···················· | ···················· | ···················· | ···················· |
| J | 3905 | hCoV-19/England/ALDP-6C43EB/2020\|EPI_ISL_556240\|20 | ···················· | ···················· | ···················· | ···················· | ···················· | ···················· |
| J | 3906 | hCoV-19/England/ALDP-6D9E5B/2020\|EPI_ISL_556241\|20 | ···················· | ···················· | ···················· | ···················· | ···················· | ···················· |
| J | 3907 | hCoV-19/England/ALDP-6CD528/2020\|EPI_ISL_556243\|20 | ···················· | ···················· | ···················· | ···················· | ···················· | ···················· |
| J | 3908 | hCoV-19/England/MILK-95D79D/2020\|EPI_ISL_556244\|20 | ···················· | ···················· | ···················· | ···················· | ···················· | ···················· |
| J | 3909 | hCoV-19/England/ALDP-6D2274/2020\|EPI_ISL_556245\|20 | ···················· | ···················· | ···················· | ···················· | ···················· | ···················· |
| J | 3910 | hCoV-19/England/ALDP-6CDE78/2020\|EPI_ISL_556246\|20 | ···················· | ···················· | ···················· | ···················· | ···················· | ···················· |
| J | 3911 | hCoV-19/England/ALDP-6D502C/2020\|EPI_ISL_556247\|20 | ···················· | ···················· | ···················· | ···················· | ···················· | ···················· |
| J | 3912 | hCoV-19/England/MILK-63E6D4/2020\|EPI_ISL_556248\|20 | ···················· | ···················· | ···················· | ···················· | ···················· | ···················· |
| J | 3913 | hCoV-19/England/ALDP-6D7A3B/2020\|EPI_ISL_556249\|20 | ···················· | ···················· | ···················· | ···················· | ···················· | ···················· |
| J | 3914 | hCoV-19/England/MILK-95D8A9/2020\|EPI_ISL_556251\|20 | ···················· | ···················· | ···················· | ···················· | ···················· | ···················· |
| J | 3915 | hCoV-19/England/ALDP-6C43DC/2020\|EPI_ISL_556252\|20 | ···················· | ···················· | ···················· | ···················· | ···················· | ···················· |
| J | 3916 | hCoV-19/England/MILK-62FD59/2020\|EPI_ISL_556253\|20 | ···················· | ···················· | ···················· | ···················· | ···················· | ···················· |
| J | 3917 | hCoV-19/England/ALDP-6C6E9D/2020\|EPI_ISL_556254\|20 | ···················· | ······A············· | ···················· | ···················· | ···················· | ···················· |
| J | 3918 | hCoV-19/England/ALDP-94EB48/2020\|EPI_ISL_556256\|20 | ···················· | ·A·················· | ···················· | ···················· | ···················· | ···················· |
| J | 3919 | hCoV-19/England/ALDP-6CCC42/2020\|EPI_ISL_556258\|20 | ···················· | ···················· | ···················· | ···················· | ···················· | ···················· |
| J | 3920 | hCoV-19/England/ALDP-6CD78C/2020\|EPI_ISL_556260\|20 | ···················· | ····A··············· | ···················· | ···················· | ···················· | ···················· |
| J | 3921 | hCoV-19/England/MILK-642628/2020\|EPI_ISL_556261\|20 | ···················· | ···················· | ···················· | ···················· | ···················· | ···················· |
| J | 3922 | hCoV-19/England/ALDP-94EB84/2020\|EPI_ISL_556262\|20 | ···················· | ···················· | ···················· | ···················· | ···················· | ···················· |
| J | 3923 | hCoV-19/England/MILK-630B88/2020\|EPI_ISL_556263\|20 | ···················· | ···················· | ···················· | ···················· | ···················· | ···················· |
| J | 3924 | hCoV-19/England/ALDP-6CD6BC/2020\|EPI_ISL_556264\|20 | ···················· | ···················· | ···················· | ···················· | ···················· | ···················· |
| J | 3925 | hCoV-19/England/ALDP-63CBF5/2020\|EPI_ISL_556265\|20 | ···················· | ···················· | ···················· | ···················· | ···················· | ···················· |
| J | 3926 | hCoV-19/England/MILK-642D7E/2020\|EPI_ISL_556266\|20 | ···················· | ···················· | ···················· | ···················· | ···················· | ···················· |
| J | 3927 | hCoV-19/England/ALDP-6CF5F9/2020\|EPI_ISL_556272\|20 | ···················· | ···················· | ···················· | ···················· | ···················· | ···················· |
| J | 3928 | hCoV-19/England/ALDP-6CD31F/2020\|EPI_ISL_556273\|20 | ···················· | ···················· | ···················· | ···················· | ···················· | ···················· |
| J | 3929 | hCoV-19/England/ALDP-6C5FF5/2020\|EPI_ISL_556274\|20 | ···················· | ···················· | ···················· | ···················· | ···················· | ···················· |
| J | 3930 | hCoV-19/England/MILK-64287D/2020\|EPI_ISL_556275\|20 | ···················· | ·A·················· | ···················· | ···················· | ···················· | ···················· |
| J | 3931 | hCoV-19/England/MILK-63F846/2020\|EPI_ISL_556276\|20 | ···················· | ···················· | ···················· | ···················· | ···················· | ···················· |
| J | 3932 | hCoV-19/England/ALDP-6D0A6F/2020\|EPI_ISL_556278\|20 | ···················· | ···················· | ···················· | ···················· | ···················· | ···················· |
| J | 3933 | hCoV-19/England/ALDP-63B7A7/2020\|EPI_ISL_556279\|20 | ···················· | ···················· | ···················· | ···················· | ···················· | ···················· |
| J | 3934 | hCoV-19/England/ALDP-95BE6D/2020\|EPI_ISL_556280\|20 | ···················· | ···················· | ···················· | ···················· | ···················· | ···················· |
| J | 3935 | hCoV-19/England/ALDP-63C6D6/2020\|EPI_ISL_556281\|20 | ···················· | ···················· | ···················· | ···················· | ···················· | ···················· |
| J | 3936 | hCoV-19/England/ALDP-94EF3D/2020\|EPI_ISL_556283\|20 | ···················· | ···················· | ···················· | ···················· | ···················· | ···················· |
| J | 3937 | hCoV-19/England/MILK-95DAEF/2020\|EPI_ISL_556285\|20 | ···················· | ···················· | ···················· | ···················· | ···················· | ···················· |
| J | 3938 | hCoV-19/England/ALDP-6DBA91/2020\|EPI_ISL_556286\|20 | ···················· | ···················· | ···················· | ···················· | ···················· | ···················· |
| J | 3939 | hCoV-19/England/ALDP-63CCA7/2020\|EPI_ISL_556288\|20 | ···················· | ···················· | ···················· | ···················· | ···················· | ···················· |
| J | 3940 | hCoV-19/England/MILK-632FD5/2020\|EPI_ISL_556289\|20 | ···················· | ···················· | ···················· | ···················· | ···················· | ···················· |
| J | 3941 | hCoV-19/England/ALDP-6C66A4/2020\|EPI_ISL_556290\|20 | ···················· | ·A·················· | ···················· | ···················· | ···················· | ···················· |
| J | 3942 | hCoV-19/England/ALDP-6CE985/2020\|EPI_ISL_556292\|20 | ···················· | ···················· | ···················· | ···················· | ···················· | ···················· |
| J | 3943 | hCoV-19/England/MILK-95D8E5/2020\|EPI_ISL_556293\|20 | ···················· | ···················· | ···················· | ···················· | ···················· | ···················· |
| J | 3944 | hCoV-19/England/MILK-642D32/2020\|EPI_ISL_556294\|20 | ···················· | ···················· | ···················· | ···················· | ···················· | ···················· |
| J | 3945 | hCoV-19/England/ALDP-6CE879/2020\|EPI_ISL_556295\|20 | ···················· | ···················· | ···················· | ···················· | ···················· | ···················· |
| J | 3946 | hCoV-19/England/ALDP-94EDCA/2020\|EPI_ISL_556297\|20 | ···················· | ···················· | ···················· | ···················· | ···················· | ···················· |
| J | 3947 | hCoV-19/England/MILK-95D7AC/2020\|EPI_ISL_556298\|20 | ···················· | ···················· | ···················· | ···················· | ···················· | ···················· |
| J | 3948 | hCoV-19/England/MILK-7512A9/2020\|EPI_ISL_557505\|20 | ···················· | ···················· | ···················· | ···················· | ···················· | ···················· |
| J | 3949 | hCoV-19/England/MILK-877A59/2020\|EPI_ISL_557506\|20 | ···················· | ···················· | ···················· | ···················· | ···················· | ···················· |
| J | 3950 | hCoV-19/England/ALDP-7440CD/2020\|EPI_ISL_557507\|20 | ···················· | ···················· | ···················· | ···················· | ···················· | ···················· |
| J | 3951 | hCoV-19/England/MILK-871433/2020\|EPI_ISL_557508\|20 | ···················· | ···················· | ···················· | ···················· | ···················· | ···················· |
| J | 3952 | hCoV-19/England/MILK-871AE6/2020\|EPI_ISL_557510\|20 | ···················· | ···················· | ···················· | ···················· | ···················· | ···················· |
| J | 3953 | hCoV-19/England/ALDP-745730/2020\|EPI_ISL_557511\|20 | ···················· | ···················· | ···················· | ···················· | ···················· | ···················· |
| J | 3954 | hCoV-19/England/MILK-74A9EA/2020\|EPI_ISL_557512\|20 | ···················· | ·············A······ | ···················· | ···················· | ···················· | ···················· |
| J | 3955 | hCoV-19/England/MILK-6F4D20/2020\|EPI_ISL_557513\|20 | ···················· | ···················· | ···················· | ···················· | ···················· | ···················· |
| J | 3956 | hCoV-19/England/MILK-86C34D/2020\|EPI_ISL_557514\|20 | ···················· | ···················· | ···················· | ···················· | ···················· | ···················· |
| J | 3957 | hCoV-19/England/MILK-85C54A/2020\|EPI_ISL_557515\|20 | ···················· | ···················· | ···················· | ···················· | ···················· | ···················· |
| J | 3958 | hCoV-19/England/MILK-865FAA/2020\|EPI_ISL_557517\|20 | ···················· | ···················· | ···················· | ···················· | ···················· | ···················· |
| J | 3959 | hCoV-19/England/ALDP-6FD880/2020\|EPI_ISL_557518\|20 | ···················· | ···················· | ···················· | ···················· | ···················· | ···················· |
| J | 3960 | hCoV-19/England/MILK-871A6E/2020\|EPI_ISL_557519\|20 | ···················· | ···················· | ···················· | ···················· | ···················· | ···················· |
| J | 3961 | hCoV-19/England/MILK-870D93/2020\|EPI_ISL_557520\|20 | ···················· | ···················· | ···················· | ···················· | ···················· | ···················· |
| J | 3962 | hCoV-19/England/MILK-6F47F5/2020\|EPI_ISL_557521\|20 | ···················· | ···················· | ···················· | ···················· | ···················· | ···················· |
| J | 3963 | hCoV-19/England/ALDP-749A33/2020\|EPI_ISL_557523\|20 | ···················· | ···················· | ···················· | ···················· | ···················· | ···················· |
| J | 3964 | hCoV-19/England/MILK-8518C4/2020\|EPI_ISL_557525\|20 | ···················· | ···················· | ···················· | ···················· | ···················· | ···················· |
| J | 3965 | hCoV-19/England/ALDP-6FC9E8/2020\|EPI_ISL_557526\|20 | ···················· | ···················· | ···················· | ···················· | ···················· | ···················· |
| J | 3966 | hCoV-19/England/ALDP-74533C/2020\|EPI_ISL_557527\|20 | ···················· | ···················· | ···················· | ···················· | ···················· | ···················· |
| J | 3967 | hCoV-19/England/MILK-85B587/2020\|EPI_ISL_557529\|20 | ···················· | ···················· | ···················· | ···················· | ···················· | ···················· |
| J | 3968 | hCoV-19/England/ALDP-6FD756/2020\|EPI_ISL_557531\|20 | ···················· | ···················· | ···················· | ···················· | ···················· | ···················· |
| J | 3969 | hCoV-19/England/MILK-6F62D4/2020\|EPI_ISL_557532\|20 | ···················· | ···················· | ···················· | ···················· | ···················· | ···················· |
| J | 3970 | hCoV-19/England/ALDP-746DD3/2020\|EPI_ISL_557533\|20 | ···················· | ···················· | ···················· | ···················· | ···················· | ···················· |
| J | 3971 | hCoV-19/England/MILK-869832/2020\|EPI_ISL_557534\|20 | ···················· | ···················· | ···················· | ···················· | ···················· | ···················· |
| J | 3972 | hCoV-19/England/MILK-74B20C/2020\|EPI_ISL_557535\|20 | ···················· | ···················· | ···················· | ···················· | ···················· | ···················· |
| J | 3973 | hCoV-19/England/MILK-74DB0F/2020\|EPI_ISL_557536\|20 | ···················· | ···················· | ···················· | ···················· | ···················· | ···················· |
| J | 3974 | hCoV-19/England/ALDP-7444FE/2020\|EPI_ISL_557537\|20 | ···················· | ···················· | ···················· | ···················· | ···················· | ···················· |
| J | 3975 | hCoV-19/England/MILK-86E6AC/2020\|EPI_ISL_557538\|20 | ···················· | ···················· | ···················· | ···················· | ···················· | ···················· |
| J | 3976 | hCoV-19/England/ALDP-74527B/2020\|EPI_ISL_557539\|20 | ···················· | ···················· | ···················· | ···················· | ···················· | ···················· |
| J | 3977 | hCoV-19/England/MILK-74C001/2020\|EPI_ISL_557540\|20 | ···················· | ···················· | ···················· | ···················· | ···················· | ···················· |
| J | 3978 | hCoV-19/England/ALDP-6FCE9E/2020\|EPI_ISL_557541\|20 | ···················· | ···················· | ···················· | ···················· | ···················· | ···················· |
| J | 3979 | hCoV-19/England/MILK-871530/2020\|EPI_ISL_557542\|20 | ···················· | ···················· | ···················· | ···················· | ···················· | ···················· |
| J | 3980 | hCoV-19/England/ALDP-748551/2020\|EPI_ISL_557543\|20 | ···················· | ···················· | ···················· | ···················· | ···················· | ···················· |

J 3981 hCoV-19/England/ALDP-6FBBE3/2020|EPI_ISL_557544|20 ···················· ···················· ····················· ···················· ···················· ····················
J 3982 hCoV-19/England/MILK-6EECA4/2020|EPI_ISL_557545|20 ···················· ···················· ····················· ···················· ···················· ····················
J 3983 hCoV-19/England/MILK-6F0B84/2020|EPI_ISL_557546|20 ···················· ···················· ····················· ···················· ···················· ····················
J 3984 hCoV-19/England/MILK-85025E/2020|EPI_ISL_557547|20 ···················· ···················· ····················· ···················· ···················· ····················
J 3985 hCoV-19/England/MILK-74B433/2020|EPI_ISL_557550|20 ···················· ···················· ····················· ···················· ···················· ····················
J 3986 hCoV-19/England/ALDP-745448/2020|EPI_ISL_557551|20 ···················· ···················· ····················· ···················· ···················· ····················
J 3987 hCoV-19/England/MILK-86F90C/2020|EPI_ISL_557552|20 ···················· ···················· ····················· ···················· ···················· ····················
J 3988 hCoV-19/England/ALDP-6FAF24/2020|EPI_ISL_557553|20 ···················· ···················· ····················· ···················· ···················· ····················
J 3989 hCoV-19/England/ALDP-6EBBD7/2020|EPI_ISL_557554|20 ···················· ···················· ····················· ···················· ···················· ····················
J 3990 hCoV-19/England/ALDP-7060F4/2020|EPI_ISL_557557|20 ···················· ···················· ····················· ···················· ···················· ····················
J 3991 hCoV-19/England/ALDP-7094F5/2020|EPI_ISL_557559|20 ···················· ···················· ····················· ···················· ···················· ····················
J 3992 hCoV-19/England/ALDP-708A12/2020|EPI_ISL_557560|20 ···················· ···················· ····················· ···················· ···················· ····················
J 3993 hCoV-19/England/ALDP-71D91D/2020|EPI_ISL_557561|20 ···················· ···················· ····················· ···················· ···················· ····················
J 3994 hCoV-19/England/ALDP-6E98D2/2020|EPI_ISL_557562|20 ···················· ···················· ····················· ···················· ···················· ····················
J 3995 hCoV-19/England/ALDP-702C7C/2020|EPI_ISL_557571|20 ···················· ···················· ····················· ···················· ···················· ····················
J 3996 hCoV-19/England/ALDP-720B8A/2020|EPI_ISL_557572|20 ···················· ···················· ····················· ···················· ···················· ····················
J 3997 hCoV-19/England/ALDP-700072/2020|EPI_ISL_557573|20 ···················· ···················· ····················· ···················· ···················· ····················
J 3998 hCoV-19/England/ALDP-704D1D/2020|EPI_ISL_557574|20 ···················· ···················· ····················· ···················· ···················· ····················
J 3999 hCoV-19/England/ALDP-71DACF/2020|EPI_ISL_557575|20 ···················· ···················· ····················· ···················· ···················· ····················
J 4000 hCoV-19/England/ALDP-702EEF/2020|EPI_ISL_557577|20 ···················· ···················· ····················· ···················· ···················· ····················
J 4001 hCoV-19/England/ALDP-6FFDDA/2020|EPI_ISL_557578|20 ···················· ···················· ····················· ···················· ···················· ····················
J 4002 hCoV-19/England/ALDP-7215D6/2020|EPI_ISL_557579|20 ···················· ···················· ····················· ···················· ···················· ····················
J 4003 hCoV-19/England/ALDP-70AB3B/2020|EPI_ISL_557580|20 ···················· ···················· ····················· ···················· ···················· ····················
J 4004 hCoV-19/England/ALDP-720416/2020|EPI_ISL_557581|20 ···················· ···················· ····················· ···················· ···················· ····················
J 4005 hCoV-19/England/ALDP-71FDA6/2020|EPI_ISL_557582|20 ···················· ···················· ····················· ···················· ···················· ····················
J 4006 hCoV-19/England/ALDP-71DCF6/2020|EPI_ISL_557583|20 ···················· ···················· ····················· ···················· ···················· ····················
J 4007 hCoV-19/England/ALDP-72158B/2020|EPI_ISL_557584|20 ···················· ···················· ····················· ···················· ···················· ····················
J 4008 hCoV-19/England/ALDP-6EAC02/2020|EPI_ISL_557586|20 ···················· ···················· ····················· ···················· ···················· ····················
J 4009 hCoV-19/England/ALDP-6EAC5D/2020|EPI_ISL_557588|20 ···················· ···················· ····················· ···················· ···················· ····················
J 4010 hCoV-19/England/ALDP-7016BB/2020|EPI_ISL_557589|20 ···················· ···················· ····················· ···················· ···················· ····················
J 4011 hCoV-19/Wales/ALDP-7058FD/2020|EPI_ISL_557590|2020 ···················· ···················· ····················· ···················· ···················· ····················
J 4012 hCoV-19/England/ALDP-70414E/2020|EPI_ISL_557591|20 ···················· ···················· ····················· ···················· ···················· ····················
J 4013 hCoV-19/England/ALDP-6E9C6D/2020|EPI_ISL_557592|20 ···················· ···················· ····················· ···················· ···················· ····················
J 4014 hCoV-19/England/ALDP-71D072/2020|EPI_ISL_557593|20 ···················· ···················· ····················· ···················· ···················· ····················
J 4015 hCoV-19/England/ALDP-70847E/2020|EPI_ISL_557594|20 ···················· ···················· ····················· ···················· ···················· ····················
J 4016 hCoV-19/England/ALDP-725156/2020|EPI_ISL_557597|20 ···················· ···················· ····················· ···················· ···················· ····················
J 4017 hCoV-19/England/ALDP-7240E1/2020|EPI_ISL_557598|20 ···················· ···················· ····················· ···················· ···················· ····················
J 4018 hCoV-19/England/ALDP-709668/2020|EPI_ISL_557599|20 ···················· ···················· ····················· ···················· ···················· ····················
J 4019 hCoV-19/England/ALDP-5BFDF5/2020|EPI_ISL_558802|20 ···················· ···················· ····················· ···················· ···················· ····················
J 4020 hCoV-19/England/ALDP-5BBF02/2020|EPI_ISL_558803|20 ···················· ···················· ····················· ···················· ···················· ····················
J 4021 hCoV-19/England/ALDP-5B465F/2020|EPI_ISL_558804|20 ···················· ···················· ····················· ···················· ···················· ····················
J 4022 hCoV-19/England/ALDP-5A48C5/2020|EPI_ISL_558805|20 ···················· ···················· ····················· ···················· ···················· ····················
J 4023 hCoV-19/England/ALDP-5A1619/2020|EPI_ISL_558806|20 ···················· ···················· ····················· ···················· ···················· ····················
J 4024 hCoV-19/England/ALDP-5C0DE2/2020|EPI_ISL_558807|20 ···················· ···················· ····················· ···················· ···················· ····················
J 4025 hCoV-19/England/ALDP-5B2B8F/2020|EPI_ISL_558808|20 ···················· ···················· ····················· ···················· ···················· ····················
J 4026 hCoV-19/England/ALDP-5AE676/2020|EPI_ISL_558809|20 ···················· ···················· ····················· ···················· ···················· ····················
J 4027 hCoV-19/Northern Ireland/ALDP-5C0CF4/2020|EPI_ISL_ ···················· ·A·················· ····················· ···················· ···················· ····················
J 4028 hCoV-19/England/ALDP-5A44EF/2020|EPI_ISL_558811|20 ···················· ···················· ····················· ···················· ···················· ····················
J 4029 hCoV-19/England/ALDP-5B9E70/2020|EPI_ISL_558815|20 ···················· ···················· ····················· ···················· ···················· ····················
J 4030 hCoV-19/England/ALDP-5A219F/2020|EPI_ISL_558816|20 ···················· ···················· ····················· ···················· ···················· ····················
J 4031 hCoV-19/England/ALDP-59F775/2020|EPI_ISL_558817|20 ···················· ···················· ····················· ···················· ···················· ····················
J 4032 hCoV-19/England/ALDP-5B23D2/2020|EPI_ISL_558818|20 ···················· ···················· ····················· ···················· ···················· ····················
J 4033 hCoV-19/England/ALDP-5A2D6E/2020|EPI_ISL_558819|20 ···················· ···················· ····················· ···················· ···················· ····················
J 4034 hCoV-19/England/ALDP-5BC217/2020|EPI_ISL_558820|20 ···················· ···················· ····················· ···················· ···················· ····················
J 4035 hCoV-19/England/ALDP-59FD28/2020|EPI_ISL_558821|20 ···················· ···················· ····················· ···················· ···················· ····················
J 4036 hCoV-19/England/ALDP-5A77B6/2020|EPI_ISL_558822|20 ···················· ···················· ····················· ···················· ···················· ····················
J 4037 hCoV-19/England/ALDP-5C4382/2020|EPI_ISL_558823|20 ···················· ···················· ····················· ···················· ···················· ····················
J 4038 hCoV-19/England/ALDP-5B6C6A/2020|EPI_ISL_558824|20 ···················· ···················· ····················· ···················· ···················· ····················
J 4039 hCoV-19/England/ALDP-5B6DA3/2020|EPI_ISL_558826|20 ···················· ···················· ····················· ···················· ···················· ····················
J 4040 hCoV-19/England/ALDP-5ABFE7/2020|EPI_ISL_558827|20 ···················· ···················· ····················· ···················· ···················· ····················
J 4041 hCoV-19/England/ALDP-5BD629/2020|EPI_ISL_558828|20 ···················· ···················· ····················· ···················· ···················· ····················
J 4042 hCoV-19/England/ALDP-5AC1FF/2020|EPI_ISL_558829|20 ···················· ···················· ····················· ···················· ···················· ····················
J 4043 hCoV-19/England/ALDP-5AC335/2020|EPI_ISL_558830|20 ···················· ···················· ····················· ···················· ···················· ····················
J 4044 hCoV-19/England/ALDP-5B53A2/2020|EPI_ISL_558831|20 ···················· ···················· ····················· ···················· ···················· ····················
J 4045 hCoV-19/England/ALDP-5BD42F/2020|EPI_ISL_558832|20 ···················· ···················· ····················· ···················· ···················· ····················
J 4046 hCoV-19/England/ALDP-5BE048/2020|EPI_ISL_558833|20 ···················· ···················· ····················· ···················· ···················· ····················
J 4047 hCoV-19/England/ALDP-5B5EFC/2020|EPI_ISL_558835|20 ···················· ···················· ····················· ···················· ···················· ····················
J 4048 hCoV-19/England/ALDP-5B142B/2020|EPI_ISL_558837|20 ···················· ···················· ····················· ···················· ···················· ····················
J 4049 hCoV-19/England/ALDP-59DB02/2020|EPI_ISL_558838|20 ···················· ···················· ····················· ···················· ···················· ····················
J 4050 hCoV-19/England/ALDP-5AFBFE/2020|EPI_ISL_558839|20 ···················· ···················· ····················· ···················· ···················· ····················
J 4051 hCoV-19/England/ALDP-5B9B79/2020|EPI_ISL_558840|20 ···················· ···················· ····················· ···················· ···················· ····················
J 4052 hCoV-19/England/ALDP-5A5A91/2020|EPI_ISL_558841|20 ···················· ···················· ····················· ···················· ···················· ····················
J 4053 hCoV-19/England/ALDP-5BF047/2020|EPI_ISL_558843|20 ···················· ···················· ····················· ···················· ···················· ····················
J 4054 hCoV-19/England/ALDP-5BF92E/2020|EPI_ISL_558844|20 ···················· ···················· ····················· ···················· ···················· ····················
J 4055 hCoV-19/England/ALDP-5B2BE9/2020|EPI_ISL_558845|20 ···················· ·A·················· ····················· ···················· ···················· ····················
J 4056 hCoV-19/England/ALDP-5AA80B/2020|EPI_ISL_558847|20 ···················· ·A·················· ····················· ···················· ···················· ····················
J 4057 hCoV-19/England/ALDP-5ADC1B/2020|EPI_ISL_558848|20 ···················· ···················· ····················· ···················· ···················· ····················
J 4058 hCoV-19/England/ALDP-5B6B12/2020|EPI_ISL_558849|20 ···················· ···················· ····················· ···················· ···················· ····················
J 4059 hCoV-19/England/ALDP-59F62D/2020|EPI_ISL_558850|20 ···················· ···················· ····················· ···················· ···················· ····················
J 4060 hCoV-19/Wales/ALDP-5C23DF/2020|EPI_ISL_558851|2020 ···················· ···················· ····················· ···················· ···················· ····················

| | | | | | | | | |
|---|---|---|---|---|---|---|---|---|
| J | 4061 | hCoV-19/England/ALDP-5B0F3A/2020\|EPI_ISL_558852\|20 | .................... | .................... | ..................... | .................... | .................... | .................... |
| J | 4062 | hCoV-19/England/ALDP-5B63FC/2020\|EPI_ISL_558853\|20 | .................... | .................... | ..................... | .................... | .................... | .................... |
| J | 4063 | hCoV-19/England/ALDP-5A4494/2020\|EPI_ISL_558854\|20 | .................... | .................... | ..................... | .................... | .................... | .................... |
| J | 4064 | hCoV-19/England/ALDP-5BCB94/2020\|EPI_ISL_558855\|20 | .................... | .................... | ..................... | .................... | .................... | .................... |
| J | 4065 | hCoV-19/England/ALDP-5B0D8B/2020\|EPI_ISL_558856\|20 | .................... | .................... | ..................... | .................... | .................... | .................... |
| J | 4066 | hCoV-19/England/ALDP-5AA698/2020\|EPI_ISL_558857\|20 | .................... | .................... | ..................... | .................... | .................... | .................... |
| J | 4067 | hCoV-19/England/ALDP-5A392D/2020\|EPI_ISL_558858\|20 | .................... | .................... | ..................... | .................... | .................... | .................... |
| J | 4068 | hCoV-19/England/ALDP-5A4142/2020\|EPI_ISL_558859\|20 | .................... | .................... | ..................... | .................... | .................... | .................... |
| J | 4069 | hCoV-19/England/ALDP-59E55E/2020\|EPI_ISL_558862\|20 | .................... | .................... | ..................... | .................... | .................... | .................... |
| J | 4070 | hCoV-19/England/ALDP-5A0BB1/2020\|EPI_ISL_558863\|20 | .................... | .................... | ..................... | .................... | .................... | .................... |
| J | 4071 | hCoV-19/England/ALDP-5B50F6/2020\|EPI_ISL_558864\|20 | .................... | .................... | ..................... | .................... | .................... | .................... |
| J | 4072 | hCoV-19/England/ALDP-5B4FDC/2020\|EPI_ISL_558865\|20 | .................... | .................... | ..................... | .................... | .................... | .................... |
| J | 4073 | hCoV-19/England/ALDP-5B81B4/2020\|EPI_ISL_558866\|20 | .................... | .................... | ..................... | .................... | .................... | .................... |
| J | 4074 | hCoV-19/England/ALDP-59FB00/2020\|EPI_ISL_558868\|20 | .................... | .................... | ..................... | .................... | .................... | .................... |
| J | 4075 | hCoV-19/England/ALDP-5A15D0/2020\|EPI_ISL_558869\|20 | .................... | .................... | ..................... | .................... | .................... | .................... |
| J | 4076 | hCoV-19/England/ALDP-5B674B/2020\|EPI_ISL_558870\|20 | .................... | .................... | ..................... | .................... | .................... | .................... |
| J | 4077 | hCoV-19/England/ALDP-5A85F7/2020\|EPI_ISL_558871\|20 | .................... | .................... | ..................... | .................... | .................... | .................... |
| J | 4078 | hCoV-19/England/ALDP-5AAD93/2020\|EPI_ISL_558873\|20 | .................... | .................... | ..................... | .................... | .................... | .................... |
| J | 4079 | hCoV-19/England/ALDP-5A2FA4/2020\|EPI_ISL_558874\|20 | .................... | .................... | ..................... | .................... | .................... | .................... |
| J | 4080 | hCoV-19/England/ALDP-5AB99E/2020\|EPI_ISL_558875\|20 | .................... | .................... | ...........T......... | .................... | .................... | .................... |
| J | 4081 | hCoV-19/England/ALDP-5A5A82/2020\|EPI_ISL_558876\|20 | .................... | .................... | ..................... | .................... | .................... | .................... |
| J | 4082 | hCoV-19/England/ALDP-5BA01F/2020\|EPI_ISL_558878\|20 | .................... | .................... | ..................... | .................... | .................... | .................... |
| J | 4083 | hCoV-19/England/ALDP-5A1E8A/2020\|EPI_ISL_558880\|20 | .................... | .................... | ..................... | .................... | .................... | .................... |
| J | 4084 | hCoV-19/England/ALDP-5C191A/2020\|EPI_ISL_558881\|20 | .................... | .................... | ..................... | .................... | .................... | .................... |
| J | 4085 | hCoV-19/England/ALDP-5A4528/2020\|EPI_ISL_558884\|20 | .................... | .................... | ..................... | .................... | .................... | .................... |
| J | 4086 | hCoV-19/England/ALDP-5B4E66/2020\|EPI_ISL_558885\|20 | .................... | .................... | ..................... | .................... | .................... | .................... |
| J | 4087 | hCoV-19/England/ALDP-5A95F6/2020\|EPI_ISL_558886\|20 | .................... | .................... | ..................... | .................... | .................... | .................... |
| J | 4088 | hCoV-19/England/ALDP-5AA7A4/2020\|EPI_ISL_558889\|20 | .................... | .A.................. | ..................... | .................... | .................... | .................... |
| J | 4089 | hCoV-19/England/ALDP-5B1959/2020\|EPI_ISL_558890\|20 | .................... | .................... | ..................... | .................... | .................... | .................... |
| J | 4090 | hCoV-19/England/ALDP-5A2A49/2020\|EPI_ISL_558892\|20 | .................... | .................... | ..................... | .................... | .................... | .................... |
| J | 4091 | hCoV-19/England/ALDP-5C42FE/2020\|EPI_ISL_558893\|20 | .................... | .................... | ..................... | .................... | .................... | .................... |
| J | 4092 | hCoV-19/England/ALDP-5C3866/2020\|EPI_ISL_558895\|20 | .................... | .................... | ..................... | .................... | .................... | .................... |
| J | 4093 | hCoV-19/England/ALDP-5B275E/2020\|EPI_ISL_558896\|20 | .................... | .................... | ..................... | .................... | .................... | .................... |
| J | 4094 | hCoV-19/England/ALDP-5BC47B/2020\|EPI_ISL_558897\|20 | .................... | .................... | ..................... | .................... | .................... | .................... |
| J | 4095 | hCoV-19/England/ALDP-5A2E10/2020\|EPI_ISL_558898\|20 | .................... | .................... | ..................... | .................... | .................... | .................... |
| J | 4096 | hCoV-19/England/ALDP-59F63C/2020\|EPI_ISL_558899\|20 | .................... | .................... | ..................... | .................... | .................... | .................... |
| J | 4097 | hCoV-19/Norway/2974/2020\|EPI_ISL_549100\|2020-07-27 | .................... | .................... | ..................... | .................... | .................... | .................... |
| J | 4098 | hCoV-19/Norway/2975/2020\|EPI_ISL_549101\|2020-08-01 | .................... | .................... | ..................... | .................... | .................... | .................... |
| J | 4099 | hCoV-19/Norway/2994/2020\|EPI_ISL_549102\|2020-07-22 | .................... | .................... | ..................... | .................... | .................... | .................... |
| J | 4100 | hCoV-19/Norway/2977/2020\|EPI_ISL_549103\|2020-07-26 | .................... | .................... | ..................... | .................... | .................... | .................... |
| J | 4101 | hCoV-19/Norway/2978/2020\|EPI_ISL_549104\|2020-07-24 | .................... | .................... | ..................... | .................... | .................... | .................... |
| J | 4102 | hCoV-19/Norway/2979/2020\|EPI_ISL_549105\|2020-07-24 | .................... | .................... | ..................... | .................... | .................... | .................... |
| J | 4103 | hCoV-19/Norway/2980/2020\|EPI_ISL_549106\|2020-07-24 | .................... | .................... | ..................... | .................... | .................... | .................... |
| J | 4104 | hCoV-19/Norway/2982/2020\|EPI_ISL_549107\|2020-07-26 | .................... | .................... | ..................... | .................... | .................... | .................... |
| J | 4105 | hCoV-19/Norway/2983/2020\|EPI_ISL_549108\|2020-07-24 | .................... | .................... | ..................... | .................... | .................... | .................... |
| J | 4106 | hCoV-19/Norway/2984/2020\|EPI_ISL_549109\|2020-07-22 | .................... | .................... | ..................... | .................... | .................... | .................... |
| J | 4107 | hCoV-19/Norway/2985/2020\|EPI_ISL_549110\|2020-07-24 | .................... | .................... | ..................... | .................... | .................... | .................... |
| J | 4108 | hCoV-19/Norway/2986/2020\|EPI_ISL_549111\|2020-07-24 | .................... | .................... | ..................... | .................... | .................... | .................... |
| J | 4109 | hCoV-19/Norway/3031/2020\|EPI_ISL_549112\|2020-08-05 | .................... | .................... | ..................... | .................... | .................... | .................... |
| J | 4110 | hCoV-19/Norway/2987/2020\|EPI_ISL_549113\|2020-07-30 | .................... | .................... | ..................... | .................... | .................... | .................... |
| J | 4111 | hCoV-19/Norway/2989/2020\|EPI_ISL_549114\|2020-07-22 | .................... | .................... | ..................... | .................... | .................... | .................... |
| J | 4112 | hCoV-19/Norway/2990/2020\|EPI_ISL_549115\|2020-07-29 | .................... | .................... | ..................... | .................... | .................... | .................... |
| J | 4113 | hCoV-19/Norway/2991/2020\|EPI_ISL_549116\|2020-07-22 | .................... | .................... | ..................... | .................... | .................... | .................... |
| J | 4114 | hCoV-19/Norway/2992/2020\|EPI_ISL_549117\|2020-07-31 | .................... | .................... | ..................... | .................... | .................... | .................... |
| J | 4115 | hCoV-19/Norway/2993/2020\|EPI_ISL_549118\|2020-07-30 | .................... | .................... | ..................... | .................... | .................... | .................... |
| J | 4116 | hCoV-19/Norway/3012/2020\|EPI_ISL_549119\|2020-08-04 | .................... | .................... | ..................... | .................... | .................... | .................... |
| J | 4117 | hCoV-19/Norway/3016/2020\|EPI_ISL_549120\|2020-08-05 | .................... | .................... | ..................... | .................... | .................... | .................... |
| J | 4118 | hCoV-19/Norway/3026/2020\|EPI_ISL_549121\|2020-08-06 | .................... | .................... | ..................... | .................... | .................... | .................... |
| J | 4119 | hCoV-19/Norway/2866/2020\|EPI_ISL_549122\|2020-07-07 | .................... | .................... | ..................... | .................... | .................... | .................... |
| J | 4120 | hCoV-19/Norway/3080/2020\|EPI_ISL_549123\|2020-08-06 | .................... | .................... | ..................... | .................... | .................... | .................... |
| J | 4121 | hCoV-19/Norway/2953/2020\|EPI_ISL_549124\|2020-07-31 | .................... | .................... | ..................... | .................... | .................... | .................... |
| J | 4122 | hCoV-19/Norway/3101/2020\|EPI_ISL_549125\|2020-08-10 | .................... | .................... | ..................... | .................... | .................... | .................... |
| J | 4123 | hCoV-19/Norway/3102/2020\|EPI_ISL_549126\|2020-08-04 | .................... | .................... | ..................... | .................... | .................... | .................... |
| J | 4124 | hCoV-19/Norway/3103/2020\|EPI_ISL_549127\|2020-08-04 | .................... | .................... | ..................... | .................... | .................... | .................... |
| J | 4125 | hCoV-19/Norway/3104/2020\|EPI_ISL_549128\|2020-08-04 | .................... | .................... | ..................... | .................... | .................... | .................... |
| J | 4126 | hCoV-19/Norway/3105/2020\|EPI_ISL_549129\|2020-08-04 | .................... | .................... | ..................... | .................... | .................... | .................... |
| J | 4127 | hCoV-19/Norway/3127/2020\|EPI_ISL_549150\|2020-08-07 | .................... | .................... | ..................... | .................... | .................... | .................... |
| J | 4128 | hCoV-19/Norway/3128/2020\|EPI_ISL_549151\|2020-08-07 | .................... | .................... | ..................... | .................... | .................... | .................... |
| J | 4129 | hCoV-19/Norway/3130/2020\|EPI_ISL_549152\|2020-08-07 | .................... | .................... | ..................... | .................... | .................... | .................... |
| J | 4130 | hCoV-19/Norway/3131/2020\|EPI_ISL_549153\|2020-08-07 | .................... | .................... | ..................... | .................... | .................... | .................... |
| J | 4131 | hCoV-19/Norway/2950/2020\|EPI_ISL_549154\|2020-07-31 | .................... | .................... | ..................... | .................... | .................... | .................... |
| J | 4132 | hCoV-19/Norway/3132/2020\|EPI_ISL_549155\|2020-08-07 | .................... | .................... | ..................... | .................... | .................... | .................... |
| J | 4133 | hCoV-19/Norway/3133/2020\|EPI_ISL_549156\|2020-08-07 | .................... | .................... | ..................... | .................... | .................... | .................... |
| J | 4134 | hCoV-19/Norway/3134/2020\|EPI_ISL_549157\|2020-08-08 | .................... | .................... | ..................... | .................... | .................... | .................... |
| J | 4135 | hCoV-19/Norway/3135/2020\|EPI_ISL_549158\|2020-08-08 | .................... | .................... | ..................... | .................... | .................... | .................... |
| J | 4136 | hCoV-19/Norway/3136/2020\|EPI_ISL_549159\|2020-08-08 | .................... | .................... | ..................... | .................... | .................... | .................... |
| J | 4137 | hCoV-19/Norway/3137/2020\|EPI_ISL_549160\|2020-08-08 | .................... | .................... | ..................... | .................... | .................... | .................... |
| J | 4138 | hCoV-19/Norway/3138/2020\|EPI_ISL_549161\|2020-08-08 | .................... | .................... | ..................... | .................... | .................... | .................... |
| J | 4139 | hCoV-19/Norway/3139/2020\|EPI_ISL_549162\|2020-08-08 | .................... | .................... | ..................... | .................... | .................... | .................... |
| J | 4140 | hCoV-19/Norway/3144/2020\|EPI_ISL_549163\|2020-08-07 | .................... | .................... | ..................... | .................... | .................... | .................... |

| | | | | | | | | |
|---|---|---|---|---|---|---|---|---|
| J | 4141 | hCoV-19/Norway/3146/2020\|EPI_ISL_549164\|2020-08-12 | ···················· | ···················· | ···················· | ···················· | ···················· | ···················· |
| J | 4142 | hCoV-19/Norway/2951/2020\|EPI_ISL_549165\|2020-07-31 | ···················· | ···················· | ···················· | ···················· | ···················· | ···················· |
| J | 4143 | hCoV-19/Norway/3147/2020\|EPI_ISL_549166\|2020-08-12 | ···················· | ···················· | ···················· | ···················· | ···················· | ···················· |
| J | 4144 | hCoV-19/Norway/3153/2020\|EPI_ISL_549167\|2020-08-17 | ···················· | ···················· | ···················· | ···················· | ···················· | ···················· |
| J | 4145 | hCoV-19/Norway/2893/2020\|EPI_ISL_549168\|2020-07-13 | ···················· | ···················· | ···················· | ···················· | ···················· | ···················· |
| J | 4146 | hCoV-19/Norway/2900/2020\|EPI_ISL_549169\|2020-07-18 | ···················· | ···················· | ···················· | ···················· | ···················· | ···················· |
| J | 4147 | hCoV-19/Norway/2902/2020\|EPI_ISL_549170\|2020-07-15 | ···················· | ···················· | ···················· | ···················· | ···················· | ···················· |
| J | 4148 | hCoV-19/Norway/2903/2020\|EPI_ISL_549171\|2020-07-15 | ···················· | ·A·················· | ···················· | ···················· | ···················· | ···················· |
| J | 4149 | hCoV-19/Norway/2904/2020\|EPI_ISL_549172\|2020-07-13 | ···················· | ·A·················· | ···················· | ···················· | ···················· | ···················· |
| J | 4150 | hCoV-19/Norway/3171/2020\|EPI_ISL_549173\|2020-06-19 | ···················· | ···················· | ···················· | ···················· | ···················· | ···················· |
| J | 4151 | hCoV-19/Norway/3173/2020\|EPI_ISL_549174\|2020-07-06 | ···················· | ·A·················· | ···················· | ···················· | ···················· | ···················· |
| J | 4152 | hCoV-19/Norway/3187/2020\|EPI_ISL_549175\|2020-08-18 | ···················· | ···················· | ···················· | ···················· | ···················· | ···················· |
| J | 4153 | hCoV-19/Sweden/RV-FOI-2/2020\|EPI_ISL_549176\|2020-0 | ···················· | ···················· | ···················· | ···················· | ···················· | ···················· |
| J | 4154 | hCoV-19/USA/FL-BPHL-1402/2020\|EPI_ISL_549184\|2020- | ···················· | ···················· | ···················· | ···················· | ···················· | ···················· |
| J | 4156 | hCoV-19/USA/FL-BPHL-1411/2020\|EPI_ISL_549193\|2020- | ···················· | ···················· | ···················· | ···················· | ···················· | ···················· |
| J | 4157 | hCoV-19/USA/FL-BPHL-1412/2020\|EPI_ISL_549194\|2020- | ···················· | ···················· | ···················· | ···················· | ···················· | ···················· |
| J | 4158 | hCoV-19/USA/FL-BPHL-1413/2020\|EPI_ISL_549195\|2020- | ···················· | ···················· | ···················· | ···················· | ···················· | ···················· |
| J | 4159 | hCoV-19/USA/FL-BPHL-1414/2020\|EPI_ISL_549196\|2020- | ···················· | ···················· | ···················· | ···················· | ···················· | ···················· |
| J | 4160 | hCoV-19/USA/FL-BPHL-1415/2020\|EPI_ISL_549197\|2020- | ···················· | ···················· | ···················· | ···················· | ···················· | ···················· |
| J | 4161 | hCoV-19/USA/FL-BPHL-1417/2020\|EPI_ISL_549199\|2020- | ···················· | ···················· | ···················· | ···················· | ···················· | ···················· |
| J | 4162 | hCoV-19/England/MILK-9A8380/2020\|EPI_ISL_550100\|20 | ···················· | ···················· | ···················· | ···················· | ···················· | ···················· |
| J | 4163 | hCoV-19/England/MILK-9A6795/2020\|EPI_ISL_550105\|20 | ···················· | ···················· | ···················· | ···················· | ···················· | ···················· |
| J | 4164 | hCoV-19/England/MILK-9A6F15/2020\|EPI_ISL_550106\|20 | ···················· | ···················· | ···················· | ···················· | ···················· | ···················· |
| J | 4165 | hCoV-19/England/MILK-9A8AF4/2020\|EPI_ISL_550107\|20 | ···················· | ·A·······A·········· | ···················· | ···················· | ···················· | ···················· |
| J | 4166 | hCoV-19/England/MILK-9A6C3C/2020\|EPI_ISL_550109\|20 | ···················· | ···················· | ···················· | ···················· | ···················· | ···················· |
| J | 4167 | hCoV-19/England/MILK-9A6971/2020\|EPI_ISL_550110\|20 | ···················· | ·A·················· | ···················· | ···················· | ···················· | ···················· |
| J | 4168 | hCoV-19/England/MILK-9A8B2E/2020\|EPI_ISL_550111\|20 | ···················· | ···················· | ···················· | ···················· | ···················· | ···················· |
| J | 4169 | hCoV-19/England/MILK-9A8AB8/2020\|EPI_ISL_550124\|20 | ···················· | ···················· | ···················· | ···················· | ···················· | ···················· |
| J | 4170 | hCoV-19/England/MILK-9A5013/2020\|EPI_ISL_550126\|20 | ···················· | ···················· | ···················· | ···················· | ···················· | ···················· |
| J | 4171 | hCoV-19/England/MILK-9A5550/2020\|EPI_ISL_550134\|20 | ···················· | ···················· | ···················· | ···················· | ···················· | ···················· |
| J | 4172 | hCoV-19/England/MILK-9A6865/2020\|EPI_ISL_550135\|20 | ···················· | ···················· | ···················· | ···················· | ···················· | ···················· |
| J | 4173 | hCoV-19/England/MILK-9A8405/2020\|EPI_ISL_550136\|20 | ···················· | ···················· | ···················· | ···················· | ···················· | ···················· |
| J | 4174 | hCoV-19/England/MILK-9A5462/2020\|EPI_ISL_550137\|20 | ···················· | ···················· | ···················· | ···················· | ···················· | ···················· |
| J | 4175 | hCoV-19/Wales/MILK-9A5347/2020\|EPI_ISL_550138\|2020 | ···················· | ···················· | ···················· | ···················· | ···················· | ···················· |
| J | 4176 | hCoV-19/England/MILK-9A6F06/2020\|EPI_ISL_550139\|20 | ···················· | ···················· | ···················· | ···················· | ···················· | ···················· |
| J | 4177 | hCoV-19/England/MILK-9A84F6/2020\|EPI_ISL_550141\|20 | ···················· | ···················· | ···················· | ···················· | ···················· | ···················· |
| J | 4178 | hCoV-19/England/MILK-9A53FC/2020\|EPI_ISL_550142\|20 | ···················· | ···················· | ···················· | ···················· | ···················· | ···················· |
| J | 4179 | hCoV-19/England/MILK-9A54AE/2020\|EPI_ISL_550143\|20 | ···················· | ···················· | ···················· | ···················· | ···················· | ···················· |
| J | 4180 | hCoV-19/Wales/MILK-9A6CC3/2020\|EPI_ISL_550145\|2020 | ···················· | ···················· | ···················· | ···················· | ···················· | ···················· |
| J | 4181 | hCoV-19/Wales/MILK-9A6CB4/2020\|EPI_ISL_550146\|2020 | ···················· | ···················· | ···················· | ···················· | ···················· | ···················· |
| J | 4182 | hCoV-19/England/MILK-9A68A1/2020\|EPI_ISL_550147\|20 | ···················· | ···················· | ···················· | ···················· | ···················· | ···················· |
| J | 4183 | hCoV-19/England/MILK-9A6EBE/2020\|EPI_ISL_550148\|20 | ···················· | ···················· | ···················· | ···················· | ···················· | ···················· |
| J | 4184 | hCoV-19/England/MILK-9A558D/2020\|EPI_ISL_550149\|20 | ···················· | ···················· | ···················· | ···················· | ···················· | ···················· |
| J | 4185 | hCoV-19/England/MILK-9A6E63/2020\|EPI_ISL_550150\|20 | ···················· | ···················· | ···················· | ···················· | ······T············· | ···················· |
| J | 4186 | hCoV-19/England/MILK-9A865A/2020\|EPI_ISL_550152\|20 | ···················· | ···················· | ···················· | ···················· | ···················· | ···················· |
| J | 4187 | hCoV-19/England/MILK-9A512F/2020\|EPI_ISL_550153\|20 | ···················· | ···················· | ···················· | ···················· | ···················· | ···················· |
| J | 4188 | hCoV-19/England/MILK-9A564E/2020\|EPI_ISL_550155\|20 | ···················· | ···················· | ···················· | ···················· | ···················· | ···················· |
| J | 4189 | hCoV-19/England/MILK-9A5365/2020\|EPI_ISL_550156\|20 | ···················· | ···················· | ···················· | ···················· | ···················· | ···················· |
| J | 4190 | hCoV-19/England/MILK-9A6A14/2020\|EPI_ISL_550157\|20 | ···················· | ···················· | ···················· | ···················· | ···················· | ···················· |
| J | 4191 | hCoV-19/England/MILK-9A846F/2020\|EPI_ISL_550158\|20 | ···················· | ···················· | ···················· | ···················· | ···················· | ···················· |
| J | 4192 | hCoV-19/Wales/MILK-9A6CD2/2020\|EPI_ISL_550159\|2020 | ···················· | ···················· | ···················· | ···················· | ···················· | ···················· |
| J | 4193 | hCoV-19/England/MILK-9A6B20/2020\|EPI_ISL_550160\|20 | ···················· | ···················· | ···················· | ···················· | ···················· | ···················· |
| J | 4194 | hCoV-19/England/MILK-9A6E36/2020\|EPI_ISL_550161\|20 | ···················· | ···················· | ···················· | ···················· | ···················· | ···················· |
| J | 4195 | hCoV-19/England/MILK-9A857B/2020\|EPI_ISL_550162\|20 | ···················· | ···················· | ···················· | ···················· | ···················· | ···················· |
| J | 4196 | hCoV-19/Wales/MILK-9A6AE7/2020\|EPI_ISL_550163\|2020 | ···················· | ···················· | ···················· | ···················· | ···················· | ···················· |
| J | 4197 | hCoV-19/England/MILK-9A8520/2020\|EPI_ISL_550164\|20 | ···················· | ·A·······A·········· | ···················· | ···················· | ···················· | ···················· |
| J | 4198 | hCoV-19/England/MILK-9A6CE1/2020\|EPI_ISL_550165\|20 | ···················· | ···················· | ···················· | ···················· | ···················· | ···················· |
| J | 4199 | hCoV-19/England/MILK-9A674A/2020\|EPI_ISL_550166\|20 | ···················· | ·········A·········· | ···················· | ···················· | ······T············· | ···················· |
| J | 4200 | hCoV-19/England/MILK-9A509B/2020\|EPI_ISL_550167\|20 | ···················· | ···················· | ···················· | ···················· | ···················· | ···················· |
| J | 4201 | hCoV-19/England/MILK-9A67C2/2020\|EPI_ISL_550169\|20 | ···················· | ···················· | ···················· | ···················· | ···················· | ···················· |
| J | 4202 | hCoV-19/England/MILK-9A6962/2020\|EPI_ISL_550170\|20 | ···················· | ···················· | ···················· | ···················· | ···················· | ···················· |
| J | 4203 | hCoV-19/England/MILK-9A5541/2020\|EPI_ISL_550172\|20 | ···················· | ···················· | ···················· | ···················· | ···················· | ···················· |
| J | 4204 | hCoV-19/England/MILK-9A6DDF/2020\|EPI_ISL_550173\|20 | ···················· | ···················· | ···················· | ···················· | ···················· | ···················· |
| J | 4205 | hCoV-19/England/MILK-9A5392/2020\|EPI_ISL_550174\|20 | ···················· | ···················· | ···················· | ···················· | ···················· | ···················· |
| J | 4206 | hCoV-19/Wales/MILK-9A6C69/2020\|EPI_ISL_550175\|2020 | ···················· | ···················· | ···················· | ···················· | ···················· | ···················· |
| J | 4207 | hCoV-19/England/MILK-9A531A/2020\|EPI_ISL_550176\|20 | ···················· | ···················· | ···················· | ···················· | ···················· | ···················· |
| J | 4208 | hCoV-19/England/MILK-9A85F3/2020\|EPI_ISL_550177\|20 | ···················· | ···················· | ···················· | ···················· | ···················· | ···················· |
| J | 4209 | hCoV-19/England/MILK-9A8CFE/2020\|EPI_ISL_550178\|20 | ···················· | ···················· | ···················· | ···················· | ···················· | ···················· |
| J | 4210 | hCoV-19/England/MILK-9A84AB/2020\|EPI_ISL_550180\|20 | ···················· | ···················· | ···················· | ···················· | ···················· | ···················· |
| J | 4211 | hCoV-19/England/MILK-9A83BD/2020\|EPI_ISL_550181\|20 | ···················· | ···················· | ···················· | ···················· | ···················· | ···················· |
| J | 4212 | hCoV-19/England/MILK-9A521D/2020\|EPI_ISL_550182\|20 | ···················· | ···················· | ···················· | ···················· | ···················· | ···················· |
| J | 4213 | hCoV-19/England/MILK-9A5189/2020\|EPI_ISL_550183\|20 | ···················· | ···················· | ···················· | ···················· | ···················· | ···················· |
| J | 4214 | hCoV-19/England/MILK-9A8D73/2020\|EPI_ISL_550184\|20 | ···················· | ···················· | ···················· | ···················· | ···················· | ···················· |
| J | 4215 | hCoV-19/Wales/MILK-9A8A30/2020\|EPI_ISL_550185\|2020 | ···················· | ···················· | ···················· | ···················· | ···················· | ···················· |
| J | 4216 | hCoV-19/Wales/MILK-9A6CA5/2020\|EPI_ISL_550186\|2020 | ···················· | ···················· | ···················· | ···················· | ···················· | ···················· |
| J | 4217 | hCoV-19/Wales/MILK-9A6B3F/2020\|EPI_ISL_550187\|2020 | ···················· | ···················· | ···················· | ···················· | ···················· | ···················· |
| J | 4218 | hCoV-19/England/MILK-9A8D0A/2020\|EPI_ISL_550188\|20 | ···················· | ···················· | ···················· | ···················· | ···················· | ···················· |
| J | 4219 | hCoV-19/England/MILK-9A85C6/2020\|EPI_ISL_550189\|20 | ···················· | ·A·················· | ···················· | ···················· | ···················· | ···A················ |
| J | 4220 | hCoV-19/England/MILK-9A8669/2020\|EPI_ISL_550190\|20 | ···················· | ···················· | ···················· | ···················· | ···················· | ···················· |
| J | 4221 | hCoV-19/England/MILK-9A567B/2020\|EPI_ISL_550191\|20 | ···················· | ···················· | ···················· | ···················· | ···················· | ···················· |

| | | | | | | | |
|---|---|---|---|---|---|---|---|
| J | 4222 hCoV-19/England/MILK-9A5523/2020\|EPI_ISL_550193\|20 | ···················· | ·A·················· | ···················· | ···················· | ···················· | ···A················ |
| J | 4223 hCoV-19/England/MILK-9A5505/2020\|EPI_ISL_550194\|20 | ···················· | ···················· | ···················· | ···················· | ···················· | ···················· |
| J | 4224 hCoV-19/England/MILK-9A8B88/2020\|EPI_ISL_550195\|20 | ···················· | ···················· | ···················· | ···················· | ···················· | ···················· |
| J | 4225 hCoV-19/England/MILK-9A50C8/2020\|EPI_ISL_550196\|20 | ···················· | ···················· | ···················· | ···················· | ···················· | ···················· |
| J | 4226 hCoV-19/England/MILK-9A864B/2020\|EPI_ISL_550197\|20 | ···················· | ···················· | ···················· | ···················· | ···················· | ···················· |
| J | 4227 hCoV-19/England/MILK-9A84D8/2020\|EPI_ISL_550198\|20 | ···················· | ···················· | ···················· | ···················· | ···················· | ···················· |
| J | 4228 hCoV-19/England/MILK-9A5408/2020\|EPI_ISL_550199\|20 | ···················· | ···················· | ···················· | ···················· | ···················· | ···················· |
| J | 4229 hCoV-19/England/ALDP-8FDC64/2020\|EPI_ISL_551401\|20 | ···················· | ···················· | ···················· | ···················· | ···················· | ···················· |
| J | 4230 hCoV-19/England/ALDP-93AA24/2020\|EPI_ISL_551403\|20 | ···················· | ···················· | ···················· | ···················· | ···················· | ···················· |
| J | 4231 hCoV-19/England/ALDP-8FB3DA/2020\|EPI_ISL_551405\|20 | ···················· | ···················· | ···················· | ···················· | ···················· | ···················· |
| J | 4232 hCoV-19/England/ALDP-8FCDEA/2020\|EPI_ISL_551407\|20 | ···················· | ···················· | ···················· | ···················· | ···················· | ···················· |
| J | 4233 hCoV-19/England/ALDP-92CFAE/2020\|EPI_ISL_551408\|20 | ···················· | ···················· | ···················· | ···················· | ···················· | ···················· |
| J | 4234 hCoV-19/England/ALDP-92D2E0/2020\|EPI_ISL_551411\|20 | ···················· | ···················· | ···················· | ···················· | ···················· | ···················· |
| J | 4235 hCoV-19/England/ALDP-8FCD53/2020\|EPI_ISL_551413\|20 | ···················· | ···················· | ···················· | ···················· | ···················· | ···················· |
| J | 4236 hCoV-19/England/ALDP-925479/2020\|EPI_ISL_551416\|20 | ···················· | ···················· | ···················· | ···················· | ···················· | ···················· |
| J | 4237 hCoV-19/England/ALDP-9055F4/2020\|EPI_ISL_551417\|20 | ···················· | ···················· | ···················· | ···················· | ···················· | ···················· |
| J | 4238 hCoV-19/England/ALDP-8FBB69/2020\|EPI_ISL_551418\|20 | ···················· | ···················· | ···················· | ···················· | ···················· | ···················· |
| J | 4239 hCoV-19/England/ALDP-8FBEE8/2020\|EPI_ISL_551419\|20 | ···················· | ···················· | ···················· | ···················· | ···················· | ···················· |
| J | 4240 hCoV-19/England/ALDP-909420/2020\|EPI_ISL_551420\|20 | ···················· | ···················· | ···················· | ···················· | ···················· | ···················· |
| J | 4241 hCoV-19/England/ALDP-92A3FF/2020\|EPI_ISL_551421\|20 | ···················· | ···················· | ···················· | ···················· | ···················· | ···················· |
| J | 4242 hCoV-19/England/ALDP-90F211/2020\|EPI_ISL_551422\|20 | ···················· | ···················· | ···················· | ···················· | ···················· | ···················· |
| J | 4243 hCoV-19/England/ALDP-938BF6/2020\|EPI_ISL_551423\|20 | ···················· | ···················· | ···················· | ···················· | ···················· | ···················· |
| J | 4244 hCoV-19/England/ALDP-92FFAB/2020\|EPI_ISL_551424\|20 | ···················· | ···················· | ···················· | ···················· | ···················· | ···················· |
| J | 4245 hCoV-19/England/ALDP-938A80/2020\|EPI_ISL_551425\|20 | ···················· | ···················· | ···················· | ···················· | ···················· | ···················· |
| J | 4246 hCoV-19/England/ALDP-936FDE/2020\|EPI_ISL_551426\|20 | ···················· | ···················· | ···················· | ···················· | ···················· | ···················· |
| J | 4247 hCoV-19/England/ALDP-93890B/2020\|EPI_ISL_551427\|20 | ···················· | ···················· | ···················· | ···················· | ···················· | ···················· |
| J | 4248 hCoV-19/England/ALDP-923B3A/2020\|EPI_ISL_551429\|20 | ···················· | ···················· | ···················· | ···················· | ···················· | ···················· |
| J | 4249 hCoV-19/England/ALDP-8FCF99/2020\|EPI_ISL_551430\|20 | ···················· | ···················· | ···················· | ···················· | ···················· | ···················· |
| J | 4250 hCoV-19/England/ALDP-924E8B/2020\|EPI_ISL_551436\|20 | ···················· | ···················· | ···················· | ···················· | ···················· | ···················· |
| J | 4251 hCoV-19/England/ALDP-92542E/2020\|EPI_ISL_551437\|20 | ···················· | ···················· | ···················· | ···················· | ···················· | ···················· |
| J | 4252 hCoV-19/England/ALDP-908CA1/2020\|EPI_ISL_551439\|20 | ···················· | ···················· | ···················· | ···················· | ···················· | ···················· |
| J | 4253 hCoV-19/England/ALDP-92D0AA/2020\|EPI_ISL_551440\|20 | ···················· | ·A·················· | ···················· | ···················· | ···················· | ···················· |
| J | 4254 hCoV-19/England/ALDP-936985/2020\|EPI_ISL_551441\|20 | ···················· | ···················· | ···················· | ···················· | ···················· | ···················· |
| J | 4255 hCoV-19/England/ALDP-927FFE/2020\|EPI_ISL_551454\|20 | ···················· | ···················· | ···················· | ···················· | ···················· | ···················· |
| J | 4256 hCoV-19/England/ALDP-923F3E/2020\|EPI_ISL_551456\|20 | ···················· | ···················· | ···················· | ···················· | ···················· | ···················· |
| J | 4257 hCoV-19/England/ALDP-938B32/2020\|EPI_ISL_551457\|20 | ···················· | ···················· | ···················· | ···················· | ···················· | ···················· |
| J | 4258 hCoV-19/England/ALDP-9081DF/2020\|EPI_ISL_551465\|20 | ···················· | ···················· | ···················· | ···················· | ···················· | ···················· |
| J | 4259 hCoV-19/England/ALDP-8FCCDE/2020\|EPI_ISL_551466\|20 | ···················· | ···················· | ···················· | ···················· | ···················· | ···················· |
| J | 4260 hCoV-19/England/ALDP-91629F/2020\|EPI_ISL_551467\|20 | ···················· | ···················· | ···················· | ···················· | ···················· | ···················· |
| J | 4261 hCoV-19/England/ALDP-90A4E3/2020\|EPI_ISL_551468\|20 | ···················· | ···················· | ···················· | ···················· | ···················· | ···················· |
| J | 4262 hCoV-19/England/ALDP-900702/2020\|EPI_ISL_551469\|20 | ···················· | ·A·················· | ···················· | ···················· | ···················· | ···················· |
| J | 4263 hCoV-19/England/ALDP-925FC3/2020\|EPI_ISL_551471\|20 | ···················· | ·A·················· | ···················· | ···················· | ···················· | ···················· |
| J | 4264 hCoV-19/England/ALDP-9057FE/2020\|EPI_ISL_551472\|20 | ···················· | ···················· | ···················· | ···················· | ···················· | ···················· |
| J | 4265 hCoV-19/England/ALDP-9042D0/2020\|EPI_ISL_551473\|20 | ···················· | ···················· | ···················· | ···················· | ···················· | ···················· |
| J | 4266 hCoV-19/England/ALDP-917A5A/2020\|EPI_ISL_551474\|20 | ···················· | ···················· | ···················· | ···················· | ···················· | ···················· |
| J | 4267 hCoV-19/England/ALDP-90812A/2020\|EPI_ISL_551475\|20 | ···················· | ···················· | ···················· | ···················· | ···················· | ···················· |
| J | 4268 hCoV-19/England/ALDP-8FD87F/2020\|EPI_ISL_551476\|20 | ···················· | ···················· | ···················· | ···················· | ···················· | ···················· |
| J | 4269 hCoV-19/England/ALDP-91A1CB/2020\|EPI_ISL_551477\|20 | ···················· | ···················· | ···················· | ···················· | ···················· | ···················· |
| J | 4270 hCoV-19/England/ALDP-93679A/2020\|EPI_ISL_551478\|20 | ···················· | ···················· | ···················· | ···················· | ···················· | ···················· |
| J | 4271 hCoV-19/England/ALDP-938BC9/2020\|EPI_ISL_551480\|20 | ···················· | ···················· | ···················· | ···················· | ···················· | ···················· |
| J | 4272 hCoV-19/England/ALDP-924EC7/2020\|EPI_ISL_551481\|20 | ···················· | ···················· | ···················· | ···················· | ···················· | ···················· |
| J | 4273 hCoV-19/England/ALDP-92650C/2020\|EPI_ISL_551482\|20 | ···················· | ···················· | ···················· | ···················· | ···················· | ···················· |
| J | 4274 hCoV-19/England/ALDP-904391/2020\|EPI_ISL_551485\|20 | ···················· | ···················· | ···················· | ···················· | ···················· | ···················· |
| J | 4275 hCoV-19/England/ALDP-92CF44/2020\|EPI_ISL_551486\|20 | ···················· | ···················· | ···················· | ···················· | ···················· | ···················· |
| J | 4276 hCoV-19/England/ALDP-91B106/2020\|EPI_ISL_551488\|20 | ···················· | ···················· | ···················· | ···················· | ···················· | ···················· |
| J | 4277 hCoV-19/England/ALDP-900A90/2020\|EPI_ISL_551489\|20 | ···················· | ···················· | ···················· | ···················· | ···················· | ···················· |
| J | 4278 hCoV-19/England/ALDP-90CEB6/2020\|EPI_ISL_551490\|20 | ···················· | ·A·················· | ···················· | ···················· | ···················· | ···················· |
| J | 4279 hCoV-19/England/ALDP-9044BC/2020\|EPI_ISL_551491\|20 | ···················· | ···················· | ···················· | ···················· | ···················· | ···················· |
| J | 4280 hCoV-19/England/ALDP-905442/2020\|EPI_ISL_551492\|20 | ···················· | ···················· | ···················· | ···················· | ···················· | ···················· |
| J | 4281 hCoV-19/England/ALDP-8FCC0B/2020\|EPI_ISL_551493\|20 | ···················· | ···················· | ···················· | ···················· | ···················· | ···················· |
| J | 4282 hCoV-19/England/ALDP-923F4D/2020\|EPI_ISL_551494\|20 | ···················· | ···················· | ···················· | ···················· | ···················· | ···················· |
| J | 4283 hCoV-19/England/ALDP-8FB53E/2020\|EPI_ISL_551495\|20 | ···················· | ···················· | ···················· | ···················· | ···················· | ···················· |
| J | 4284 hCoV-19/England/ALDP-904856/2020\|EPI_ISL_551497\|20 | ···················· | ···················· | ···················· | ···················· | ···················· | ···················· |
| J | 4285 hCoV-19/England/ALDP-92D022/2020\|EPI_ISL_551498\|20 | ···················· | ···················· | ···················· | ···················· | ···················· | ···················· |
| J | 4286 hCoV-19/England/ALDP-8FD8F7/2020\|EPI_ISL_551499\|20 | ···················· | ···················· | ···················· | ···················· | ···················· | ···················· |
| J | 4287 hCoV-19/England/MILK-975823/2020\|EPI_ISL_552700\|20 | ···················· | ···················· | ···················· | ···················· | ···················· | ···················· |
| J | 4288 hCoV-19/England/MILK-9751DD/2020\|EPI_ISL_552701\|20 | ···················· | ···················· | ···················· | ···················· | ···················· | ···················· |
| J | 4289 hCoV-19/England/MILK-9750D0/2020\|EPI_ISL_552702\|20 | ···················· | ···················· | ···················· | ···················· | ···················· | ···················· |
| J | 4290 hCoV-19/England/ALDP-982DCA/2020\|EPI_ISL_552703\|20 | ···················· | ···················· | ···················· | ···················· | ···················· | ···················· |
| J | 4291 hCoV-19/England/MILK-9755FF/2020\|EPI_ISL_552704\|20 | ···················· | ···················· | ···················· | ···················· | ···················· | ···················· |
| J | 4292 hCoV-19/England/MILK-97597B/2020\|EPI_ISL_552705\|20 | ···················· | ···················· | ···················· | ···················· | ···················· | ···················· |
| J | 4293 hCoV-19/England/ALDP-982B66/2020\|EPI_ISL_552706\|20 | ···················· | ·A·················· | ···················· | ···················· | ···················· | ···················· |
| J | 4294 hCoV-19/England/ALDP-982D60/2020\|EPI_ISL_552707\|20 | ···················· | ···················· | ···················· | ···················· | ···················· | ···················· |
| J | 4295 hCoV-19/England/MILK-975AA5/2020\|EPI_ISL_552708\|20 | ···················· | ···················· | ···················· | ···················· | ···················· | ···················· |
| J | 4296 hCoV-19/England/MILK-9757CC/2020\|EPI_ISL_552709\|20 | ···················· | ·······A············ | ···················· | ···················· | ·····T·············· | ···················· |
| J | 4297 hCoV-19/England/MILK-9752F8/2020\|EPI_ISL_552710\|20 | ···················· | ···················· | ···················· | ···················· | ·····T·············· | ···················· |
| J | 4298 hCoV-19/England/ALDP-982BEE/2020\|EPI_ISL_552712\|20 | ···················· | ···················· | ···················· | ···················· | ···················· | ···················· |
| J | 4299 hCoV-19/England/ALDP-98297B/2020\|EPI_ISL_552713\|20 | ···················· | ···················· | ···················· | ···················· | ···················· | ···················· |
| J | 4300 hCoV-19/England/MILK-974F4D/2020\|EPI_ISL_552714\|20 | ···················· | ···················· | ···················· | ···················· | ···················· | ···················· |
| J | 4301 hCoV-19/England/MILK-975C54/2020\|EPI_ISL_552715\|20 | ···················· | ···················· | ···················· | ···················· | ···················· | ···················· |

| | | | | | | | | | |
|---|---|---|---|---|---|---|---|---|---|
| J | 4302 | hCoV-19/England/MILK-976039/2020\|EPI_ISL_552718\|20 | ···················· | ···················· | ····················· | ···················· | ···················· | ···················· | |
| J | 4303 | hCoV-19/England/ALDP-981C46/2020\|EPI_ISL_552721\|20 | ···················· | ···················· | ····················· | ···················· | ···················· | ···················· | |
| J | 4304 | hCoV-19/England/ALDP-981CEC/2020\|EPI_ISL_552723\|20 | ···················· | ···················· | ····················· | ···················· | ···················· | ···················· | |
| J | 4305 | hCoV-19/England/ALDP-981F10/2020\|EPI_ISL_552724\|20 | ···················· | ·A·················· | ····················· | ···················· | ···················· | ···················· | |
| J | 4306 | hCoV-19/Wales/MILK-975D24/2020\|EPI_ISL_552725\|2020 | ···················· | ···················· | ····················· | ···················· | ···················· | ···················· | |
| J | 4307 | hCoV-19/England/ALDP-981FE3/2020\|EPI_ISL_552726\|20 | ···················· | ···················· | ····················· | ···················· | ···················· | ···················· | |
| J | 4308 | hCoV-19/England/MILK-975D42/2020\|EPI_ISL_552727\|20 | ···················· | ···················· | ····················· | ···················· | ···················· | ···················· | |
| J | 4309 | hCoV-19/England/ALDP-981D07/2020\|EPI_ISL_552728\|20 | ···················· | ···················· | ····················· | ···················· | ···················· | ···················· | |
| J | 4310 | hCoV-19/England/ALDP-981CFB/2020\|EPI_ISL_552731\|20 | ···················· | ·A·················· | ····················· | ···················· | ···················· | ···················· | |
| J | 4311 | hCoV-19/England/ALDP-982058/2020\|EPI_ISL_552732\|20 | ···················· | ···················· | ····················· | ···················· | ···················· | ···················· | |
| J | 4312 | hCoV-19/England/MILK-9760EE/2020\|EPI_ISL_552733\|20 | ···················· | ···················· | ····················· | ···················· | ···················· | ···················· | |
| J | 4313 | hCoV-19/England/ALDP-981E22/2020\|EPI_ISL_552735\|20 | ···················· | ···················· | ····················· | ···················· | ···················· | ···················· | |
| J | 4314 | hCoV-19/England/MILK-975D9D/2020\|EPI_ISL_552736\|20 | ···················· | ···················· | ···········T········· | ···················· | ···················· | ···················· | |
| J | 4315 | hCoV-19/England/MILK-975E9A/2020\|EPI_ISL_552738\|20 | ···················· | ···················· | ····················· | ···················· | ···················· | ···················· | |
| J | 4316 | hCoV-19/Wales/MILK-975CAF/2020\|EPI_ISL_552740\|2020 | ···················· | ···················· | ····················· | ···················· | ···················· | ···················· | |
| J | 4317 | hCoV-19/England/ALDP-981F7A/2020\|EPI_ISL_552741\|20 | ···················· | ···················· | ····················· | ···················· | ···················· | ···················· | |
| J | 4318 | hCoV-19/England/ALDP-981C0A/2020\|EPI_ISL_552742\|20 | ···················· | ···················· | ····················· | ···················· | ···················· | ···················· | |
| J | 4319 | hCoV-19/England/ALDP-981CDD/2020\|EPI_ISL_552746\|20 | ···················· | ···················· | ····················· | ···················· | ···················· | ···················· | |
| J | 4320 | hCoV-19/England/MILK-975FB5/2020\|EPI_ISL_552747\|20 | ···················· | ···················· | ····················· | ···················· | ···················· | ···················· | |
| J | 4321 | hCoV-19/Wales/MILK-975CBE/2020\|EPI_ISL_552752\|2020 | ···················· | ···················· | ····················· | ···················· | ···················· | ···················· | |
| J | 4322 | hCoV-19/England/MILK-976118/2020\|EPI_ISL_552753\|20 | ···················· | ···················· | ····················· | ···················· | ······T············· | ···················· | |
| J | 4323 | hCoV-19/England/MILK-976743/2020\|EPI_ISL_552756\|20 | ···················· | ···················· | ····················· | ···················· | ···················· | ···················· | |
| J | 4324 | hCoV-19/Wales/MILK-975CFA/2020\|EPI_ISL_552759\|2020 | ···················· | ···················· | ····················· | ···················· | ···················· | ···················· | |
| J | 4325 | hCoV-19/England/ALDP-981FB6/2020\|EPI_ISL_552760\|20 | ···················· | ···················· | ····················· | ···················· | ···················· | ···················· | |
| J | 4326 | hCoV-19/England/MILK-975EB8/2020\|EPI_ISL_552767\|20 | ···················· | ···················· | ····················· | ···················· | ···················· | ···················· | |
| J | 4327 | hCoV-19/Wales/MILK-97685F/2020\|EPI_ISL_552769\|2020 | ···················· | ···················· | ····················· | ···················· | ···················· | ···················· | |
| J | 4328 | hCoV-19/England/MILK-975D33/2020\|EPI_ISL_552772\|20 | ···················· | ···················· | ····················· | ···················· | ···················· | ···················· | |
| J | 4329 | hCoV-19/England/ALDP-9820FE/2020\|EPI_ISL_552773\|20 | ···················· | ···················· | ····················· | ···················· | ···················· | ···················· | |
| J | 4330 | hCoV-19/England/ALDP-981D8F/2020\|EPI_ISL_552785\|20 | ···················· | ···················· | ····················· | ···················· | ···················· | ···················· | |
| J | 4331 | hCoV-19/England/MILK-97694D/2020\|EPI_ISL_552788\|20 | ···················· | ···················· | ····················· | ···················· | ···················· | ···················· | |
| J | 4332 | hCoV-19/England/ALDP-981DAD/2020\|EPI_ISL_552796\|20 | ···················· | ·A·················· | ····················· | ···················· | ···················· | ···················· | |
| J | 4333 | hCoV-19/England/ALDP-935C6F/2020\|EPI_ISL_552797\|20 | ···················· | ···················· | ····················· | ···················· | ···················· | ···················· | |
| J | 4334 | hCoV-19/England/ALDP-913E43/2020\|EPI_ISL_552798\|20 | ···················· | ···················· | ····················· | ···················· | ···················· | ···················· | |
| J | 4335 | hCoV-19/England/MILK-93A0E6/2020\|EPI_ISL_552799\|20 | ···················· | ···················· | ····················· | ···················· | ···················· | ···················· | |
| J | 4336 | hCoV-19/England/ALDP-5A39E1/2020\|EPI_ISL_558900\|20 | ···················· | ···················· | ····················· | ···················· | ···················· | ···················· | |
| J | 4337 | hCoV-19/England/ALDP-519D53/2020\|EPI_ISL_558901\|20 | ···················· | ···················· | ····················· | ···················· | ···················· | ··················C· | 1 |
| J | 4338 | hCoV-19/England/ALDP-52698C/2020\|EPI_ISL_558902\|20 | ···················· | ···················· | ····················· | ···················· | ···················· | ···················· | |
| J | 4339 | hCoV-19/England/ALDP-51E064/2020\|EPI_ISL_558904\|20 | ···················· | ···················· | ····················· | ···················· | ···················· | ···················· | |
| J | 4340 | hCoV-19/England/ALDP-524942/2020\|EPI_ISL_558905\|20 | ···················· | ···················· | ····················· | ···················· | ···················· | ···················· | |
| J | 4341 | hCoV-19/England/ALDP-52A3E4/2020\|EPI_ISL_558906\|20 | ···················· | ···················· | ····················· | ···················· | ···················· | ···················· | |
| J | 4342 | hCoV-19/England/ALDP-5200E7/2020\|EPI_ISL_558907\|20 | ···················· | ···················· | ····················· | ···················· | ···················· | ···················· | |
| J | 4343 | hCoV-19/England/ALDP-51F5CE/2020\|EPI_ISL_558908\|20 | ···················· | ···················· | ····················· | ···················· | ···················· | ···················· | |
| J | 4344 | hCoV-19/Wales/ALDP-52C063/2020\|EPI_ISL_558909\|2020 | ···················· | ···················· | ····················· | ···················· | ···················· | ···················· | |
| J | 4345 | hCoV-19/England/ALDP-520199/2020\|EPI_ISL_558910\|20 | ···················· | ···················· | ····················· | ···················· | ···················· | ···················· | |
| J | 4346 | hCoV-19/England/ALDP-521C3D/2020\|EPI_ISL_558911\|20 | ···················· | ···················· | ····················· | ···················· | ···················· | ···················· | |
| J | 4347 | hCoV-19/England/ALDP-52B398/2020\|EPI_ISL_558912\|20 | ···················· | ···················· | ····················· | ···················· | ···················· | ···················· | |
| J | 4348 | hCoV-19/England/MILK-50FCEA/2020\|EPI_ISL_558914\|20 | ···················· | ···················· | ····················· | ···················· | ···················· | ···················· | |
| J | 4349 | hCoV-19/England/ALDP-5275F0/2020\|EPI_ISL_558917\|20 | ···················· | ···················· | ····················· | ···················· | ···················· | ···················· | |
| J | 4350 | hCoV-19/England/ALDP-522FAC/2020\|EPI_ISL_558918\|20 | ···················· | ·A·················· | ····················· | ···················· | ···················· | ···················· | |
| J | 4351 | hCoV-19/England/MILK-51664C/2020\|EPI_ISL_558920\|20 | ···················· | ···················· | ····················· | ···················· | ···················· | ···················· | |
| J | 4352 | hCoV-19/England/ALDP-52B9E1/2020\|EPI_ISL_558922\|20 | ···················· | ···················· | ····················· | ···················· | ···················· | ···················· | |
| J | 4353 | hCoV-19/Norway/3106/2020\|EPI_ISL_549130\|2020-08-04 | ···················· | ···················· | ····················· | ···················· | ···················· | ···················· | |
| J | 4354 | hCoV-19/Norway/3108/2020\|EPI_ISL_549131\|2020-08-05 | ···················· | ···················· | ····················· | ···················· | ···················· | ···················· | |
| J | 4355 | hCoV-19/Norway/2889/2020\|EPI_ISL_549132\|2020-07-14 | ···················· | ···················· | ····················· | ···················· | ···················· | ···················· | |
| J | 4356 | hCoV-19/Norway/3110/2020\|EPI_ISL_549133\|2020-08-05 | ···················· | ···················· | ····················· | ···················· | ···················· | ···················· | |
| J | 4357 | hCoV-19/Norway/3111/2020\|EPI_ISL_549134\|2020-08-05 | ···················· | ···················· | ····················· | ···················· | ···················· | ···················· | |
| J | 4358 | hCoV-19/Norway/3112/2020\|EPI_ISL_549135\|2020-08-05 | ···················· | ···················· | ····················· | ···················· | ···················· | ···················· | |
| J | 4359 | hCoV-19/Norway/3113/2020\|EPI_ISL_549136\|2020-08-05 | ···················· | ···················· | ····················· | ···················· | ···················· | ···················· | |
| J | 4360 | hCoV-19/Norway/3114/2020\|EPI_ISL_549137\|2020-08-06 | ···················· | ···················· | ····················· | ···················· | ···················· | ···················· | |
| J | 4361 | hCoV-19/Norway/3115/2020\|EPI_ISL_549138\|2020-08-06 | ···················· | ···················· | ····················· | ···················· | ···················· | ···················· | |
| J | 4362 | hCoV-19/Norway/3116/2020\|EPI_ISL_549139\|2020-08-06 | ···················· | ···················· | ····················· | ···················· | ···················· | ···················· | |
| J | 4363 | hCoV-19/England/MILK-99CC1B/2020\|EPI_ISL_549815\|20 | ···················· | ···················· | ····················· | ···················· | ···················· | ···················· | |
| J | 4364 | hCoV-19/England/MILK-99CD45/2020\|EPI_ISL_549816\|20 | ···················· | ···················· | ····················· | ···················· | ···················· | ···················· | |
| J | 4365 | hCoV-19/England/MILK-99D5B5/2020\|EPI_ISL_549817\|20 | ···················· | ······A············· | ····················· | ···················· | ···················· | ···················· | |
| J | 4366 | hCoV-19/England/MILK-99CD09/2020\|EPI_ISL_549818\|20 | ···················· | ···················· | ····················· | ···················· | ···················· | ···················· | |
| J | 4367 | hCoV-19/England/MILK-99CE8E/2020\|EPI_ISL_549819\|20 | ···················· | ···················· | ····················· | ···················· | ···················· | ···················· | |
| J | 4368 | hCoV-19/England/MILK-99D4E5/2020\|EPI_ISL_549820\|20 | ···················· | ···················· | ····················· | ···················· | ···················· | ···················· | |
| J | 4369 | hCoV-19/Wales/MILK-99D01E/2020\|EPI_ISL_549821\|2020 | ···················· | ···················· | ····················· | ···················· | ···················· | ···················· | |
| J | 4370 | hCoV-19/England/MILK-99CB78/2020\|EPI_ISL_549822\|20 | ···················· | ···················· | ····················· | ···················· | ···················· | ···················· | |
| J | 4371 | hCoV-19/England/MILK-99D02D/2020\|EPI_ISL_549823\|20 | ···················· | ···················· | ····················· | ···················· | ···················· | ···················· | |
| J | 4372 | hCoV-19/England/MILK-99D11B/2020\|EPI_ISL_549825\|20 | ···················· | ···················· | ····················· | ···················· | ···················· | ···················· | |
| J | 4373 | hCoV-19/England/MILK-99D069/2020\|EPI_ISL_549826\|20 | ···················· | ···················· | ····················· | ···················· | ···················· | ···················· | |
| J | 4374 | hCoV-19/England/MILK-99D078/2020\|EPI_ISL_549828\|20 | ···················· | ···················· | ····················· | ···················· | ···················· | ···················· | |
| J | 4375 | hCoV-19/England/MILK-99D281/2020\|EPI_ISL_549830\|20 | ···················· | ···················· | ····················· | ···················· | ···················· | ···················· | |
| J | 4376 | hCoV-19/England/MILK-99CE15/2020\|EPI_ISL_549831\|20 | ···················· | ···················· | ····················· | ···················· | ···················· | ···················· | |
| J | 4377 | hCoV-19/England/MILK-99D597/2020\|EPI_ISL_549833\|20 | ···················· | ···················· | ····················· | ···················· | ···················· | ···················· | |
| J | 4378 | hCoV-19/England/MILK-99CCC0/2020\|EPI_ISL_549834\|20 | ···················· | ···················· | ····················· | ···················· | ···················· | ···················· | |
| J | 4379 | hCoV-19/England/MILK-99CE24/2020\|EPI_ISL_549835\|20 | ···················· | ···················· | ····················· | ···················· | ···················· | ···················· | |
| J | 4380 | hCoV-19/England/MILK-99CE51/2020\|EPI_ISL_549837\|20 | ···················· | ···················· | ····················· | ···················· | ···················· | ···················· | |
| J | 4381 | hCoV-19/England/MILK-99CCFD/2020\|EPI_ISL_549838\|20 | ···················· | ···················· | ····················· | ···················· | ···················· | ···················· | |

| | | | | | | | | |
|---|---|---|---|---|---|---|---|---|
| J | 4382 | hCoV-19/England/MILK-99CCDF/2020\|EPI_ISL_549839\|20 | ···················· | ···················· | ····················· | ···················· | ···················· | ···················· |
| J | 4383 | hCoV-19/England/MILK-99D0F0/2020\|EPI_ISL_549840\|20 | ···················· | ···················· | ····················· | ···················· | ···················· | ···················· |
| J | 4384 | hCoV-19/England/MILK-99CEF7/2020\|EPI_ISL_549841\|20 | ···················· | ·A·················· | ····················· | ···················· | ···················· | ···A················ |
| J | 4385 | hCoV-19/England/MILK-99CD36/2020\|EPI_ISL_549842\|20 | ···················· | ···················· | ····················· | ···················· | ···················· | ···················· |
| J | 4386 | hCoV-19/England/MILK-99CB69/2020\|EPI_ISL_549843\|20 | ···················· | ···················· | ····················· | ···················· | ···················· | ···················· |
| J | 4387 | hCoV-19/England/MILK-99CEE8/2020\|EPI_ISL_549844\|20 | ···················· | ·A·················· | ····················· | ···················· | ···················· | ···A················ |
| J | 4388 | hCoV-19/England/MILK-99D5E2/2020\|EPI_ISL_549845\|20 | ···················· | ···················· | ····················· | ···················· | ···················· | ···················· |
| J | 4389 | hCoV-19/England/MILK-99CF7C/2020\|EPI_ISL_549846\|20 | ···················· | ···················· | ····················· | ···················· | ···················· | ···················· |
| J | 4390 | hCoV-19/England/MILK-99CEBB/2020\|EPI_ISL_549847\|20 | ···················· | ···················· | ····················· | ···················· | ···················· | ···················· |
| J | 4391 | hCoV-19/England/MILK-99CC48/2020\|EPI_ISL_549848\|20 | ···················· | ···················· | ····················· | ···················· | ···················· | ···················· |
| J | 4392 | hCoV-19/England/MILK-99CD54/2020\|EPI_ISL_549849\|20 | ···················· | ···················· | ····················· | ···················· | ···················· | ···················· |
| J | 4393 | hCoV-19/England/MILK-99CFE5/2020\|EPI_ISL_549850\|20 | ···················· | ···················· | ····················· | ···················· | ···················· | ···················· |
| J | 4394 | hCoV-19/England/MILK-99CC75/2020\|EPI_ISL_549851\|20 | ···················· | ···················· | ····················· | ···················· | ···················· | ···················· |
| J | 4395 | hCoV-19/England/MILK-99D2FA/2020\|EPI_ISL_549852\|20 | ···················· | ···················· | ····················· | ···················· | ···················· | ···················· |
| J | 4396 | hCoV-19/England/MILK-99CCA2/2020\|EPI_ISL_549853\|20 | ···················· | ···················· | ····················· | ···················· | ···················· | ···················· |
| J | 4397 | hCoV-19/England/MILK-99CFF4/2020\|EPI_ISL_549854\|20 | ···················· | ···················· | ····················· | ···················· | ···················· | ···················· |
| J | 4398 | hCoV-19/England/MILK-99D272/2020\|EPI_ISL_549855\|20 | ···················· | ···················· | ····················· | ···················· | ···················· | ···················· |
| J | 4399 | hCoV-19/England/MILK-99CB87/2020\|EPI_ISL_549856\|20 | ···················· | ···················· | ····················· | ···················· | ···················· | ···················· |
| J | 4400 | hCoV-19/England/MILK-99D500/2020\|EPI_ISL_549857\|20 | ···················· | ···················· | ····················· | ···················· | ···················· | ···················· |
| J | 4401 | hCoV-19/England/MILK-99D588/2020\|EPI_ISL_549858\|20 | ···················· | ····A··············· | ····················· | ···················· | ···················· | ···················· |
| J | 4402 | hCoV-19/England/MILK-99CE9D/2020\|EPI_ISL_549859\|20 | ···················· | ···················· | ····················· | ···················· | ···················· | ···················· |
| J | 4403 | hCoV-19/England/MILK-99D0E1/2020\|EPI_ISL_549860\|20 | ···················· | ···················· | ····················· | ···················· | ···················· | ···················· |
| J | 4404 | hCoV-19/England/MILK-99D6D0/2020\|EPI_ISL_549862\|20 | ···················· | ···················· | ····················· | ···················· | ···················· | ···················· |
| J | 4405 | hCoV-19/Wales/MILK-99D04B/2020\|EPI_ISL_549863\|2020 | ···················· | ···················· | ····················· | ···················· | ···················· | ···················· |
| J | 4406 | hCoV-19/England/MILK-99CB5A/2020\|EPI_ISL_549864\|20 | ···················· | ···················· | ····················· | ···················· | ···················· | ···················· |
| J | 4407 | hCoV-19/England/MILK-99D38E/2020\|EPI_ISL_549865\|20 | ···················· | ···················· | ····················· | ···················· | ···················· | ···················· |
| J | 4408 | hCoV-19/England/MILK-99D403/2020\|EPI_ISL_549866\|20 | ···················· | ···················· | ····················· | ···················· | ···················· | ···················· |
| J | 4409 | hCoV-19/England/MILK-99D62B/2020\|EPI_ISL_549867\|20 | ···················· | ·········A·········· | ····················· | ···················· | ······T············· | ···················· |
| J | 4410 | hCoV-19/England/MILK-99D096/2020\|EPI_ISL_549868\|20 | ···················· | ···················· | ····················· | ···················· | ···················· | ···················· |
| J | 4411 | hCoV-19/England/MILK-99CB96/2020\|EPI_ISL_549869\|20 | ···················· | ···················· | ····················· | ···················· | ···················· | ···················· |
| J | 4412 | hCoV-19/England/MILK-99D37F/2020\|EPI_ISL_549870\|20 | ···················· | ·A·······A·········· | ····················· | ···················· | ···················· | ···················· |
| J | 4413 | hCoV-19/England/MILK-99CBC3/2020\|EPI_ISL_549871\|20 | ···················· | ···················· | ····················· | ···················· | ······T············· | ···················· |
| J | 4414 | hCoV-19/England/MILK-99D56A/2020\|EPI_ISL_549872\|20 | ···················· | ···················· | ····················· | ···················· | ···················· | ···················· |
| J | 4415 | hCoV-19/England/MILK-99D764/2020\|EPI_ISL_549874\|20 | ···················· | ···················· | ····················· | ···················· | ···················· | ···················· |
| J | 4416 | hCoV-19/England/MILK-99D3CA/2020\|EPI_ISL_549875\|20 | ···················· | ···················· | ····················· | ···················· | ···················· | ···················· |
| J | 4417 | hCoV-19/England/MILK-99D3AC/2020\|EPI_ISL_549876\|20 | ···················· | ···················· | ····················· | ···················· | ···················· | ···················· |
| J | 4418 | hCoV-19/England/MILK-99D03C/2020\|EPI_ISL_549877\|20 | ···················· | ···················· | ····················· | ···················· | ···················· | ···················· |
| J | 4419 | hCoV-19/England/MILK-99CD63/2020\|EPI_ISL_549878\|20 | ···················· | ···················· | ····················· | ···················· | ···················· | ···················· |
| J | 4420 | hCoV-19/England/MILK-99D48B/2020\|EPI_ISL_549879\|20 | ···················· | ···················· | ····················· | ···················· | ···················· | ···················· |
| J | 4421 | hCoV-19/England/MILK-99D3BB/2020\|EPI_ISL_549880\|20 | ···················· | ···················· | ····················· | ···················· | ···················· | ···················· |
| J | 4422 | hCoV-19/England/MILK-99D360/2020\|EPI_ISL_549881\|20 | ···················· | ···················· | ····················· | ···················· | ···················· | ···················· |
| J | 4423 | hCoV-19/England/MILK-99D685/2020\|EPI_ISL_549882\|20 | ···················· | ···················· | ····················· | ···················· | ···················· | ···················· |
| J | 4424 | hCoV-19/England/MILK-99D4B8/2020\|EPI_ISL_549883\|20 | ···················· | ···················· | ····················· | ···················· | ···················· | ···················· |
| J | 4425 | hCoV-19/England/MILK-99CBE1/2020\|EPI_ISL_549886\|20 | ···················· | ···················· | ····················· | ···················· | ···················· | ···················· |
| J | 4426 | hCoV-19/England/MILK-99D773/2020\|EPI_ISL_549887\|20 | ···················· | ···················· | ····················· | ···················· | ···················· | ···················· |
| J | 4427 | hCoV-19/England/MILK-99D63A/2020\|EPI_ISL_549888\|20 | ···················· | ···················· | ····················· | ···················· | ···················· | ···················· |
| J | 4428 | hCoV-19/England/MILK-99D46D/2020\|EPI_ISL_549889\|20 | ···················· | ···················· | ····················· | ···················· | ···················· | ···················· |
| J | 4429 | hCoV-19/England/MILK-99CC57/2020\|EPI_ISL_549890\|20 | ···················· | ·A·················· | ····················· | ···················· | ···················· | ···················· |
| J | 4430 | hCoV-19/England/MILK-99CD90/2020\|EPI_ISL_549891\|20 | ···················· | ···················· | ····················· | ···················· | ···················· | ···················· |
| J | 4431 | hCoV-19/England/MILK-99D306/2020\|EPI_ISL_549892\|20 | ···················· | ···················· | ···········T········· | ···················· | ···················· | ···················· |
| J | 4432 | hCoV-19/England/MILK-99CF8B/2020\|EPI_ISL_549893\|20 | ···················· | ···················· | ····················· | ···················· | ···················· | ···················· |
| J | 4433 | hCoV-19/England/MILK-99D44F/2020\|EPI_ISL_549894\|20 | ···················· | ···················· | ····················· | ···················· | ······T············· | ···················· |
| J | 4434 | hCoV-19/England/MILK-99CD72/2020\|EPI_ISL_549895\|20 | ···················· | ···················· | ····················· | ···················· | ···················· | ···················· |
| J | 4435 | hCoV-19/England/MILK-99D00F/2020\|EPI_ISL_549896\|20 | ···················· | ···················· | ····················· | ···················· | ···················· | ···················· |
| J | 4436 | hCoV-19/England/MILK-99D49A/2020\|EPI_ISL_549897\|20 | ···················· | ···················· | ····················· | ···················· | ···················· | ···················· |
| J | 4437 | hCoV-19/England/MILK-99CC2A/2020\|EPI_ISL_549898\|20 | ···················· | ···················· | ····················· | ···················· | ···················· | ···················· |
| J | 4438 | hCoV-19/England/MILK-99CEAC/2020\|EPI_ISL_549899\|20 | ···················· | ···················· | ····················· | ···················· | ···················· | ···················· |
| J | 4439 | hCoV-19/England/MILK-9A8B79/2020\|EPI_ISL_550101\|20 | ···················· | ···················· | ····················· | ···················· | ···················· | ···················· |
| J | 4440 | hCoV-19/England/MILK-9A6A9C/2020\|EPI_ISL_550102\|20 | ···················· | ···················· | ····················· | ···················· | ···················· | ···················· |
| J | 4441 | hCoV-19/England/MILK-9A6935/2020\|EPI_ISL_550103\|20 | ···················· | ···················· | ····················· | ···················· | ···················· | ···················· |
| J | 4442 | hCoV-19/England/MILK-9A6E09/2020\|EPI_ISL_550104\|20 | ···················· | ···················· | ····················· | ···················· | ···················· | ···················· |
| J | 4443 | hCoV-19/England/MILK-9A8C67/2020\|EPI_ISL_550108\|20 | ···················· | ···················· | ····················· | ···················· | ···················· | ···················· |
| J | 4444 | hCoV-19/England/MILK-9A8362/2020\|EPI_ISL_550112\|20 | ···················· | ···················· | ····················· | ···················· | ···················· | ···················· |
| J | 4445 | hCoV-19/England/MILK-9A5787/2020\|EPI_ISL_550113\|20 | ···················· | ···················· | ····················· | ···················· | ···················· | ···················· |
| J | 4446 | hCoV-19/England/MILK-9A671D/2020\|EPI_ISL_550114\|20 | ···················· | ···················· | ····················· | ···················· | ···················· | ···················· |
| J | 4447 | hCoV-19/England/MILK-9A5259/2020\|EPI_ISL_550115\|20 | ···················· | ···················· | ····················· | ···················· | ···················· | ···················· |
| J | 4448 | hCoV-19/England/MILK-9A6777/2020\|EPI_ISL_550116\|20 | ···················· | ···················· | ····················· | ···················· | ···················· | ···················· |
| J | 4449 | hCoV-19/England/MILK-9A68FC/2020\|EPI_ISL_550117\|20 | ···················· | ···················· | ····················· | ···················· | ···················· | ···················· |
| J | 4450 | hCoV-19/England/MILK-9A855D/2020\|EPI_ISL_550118\|20 | ···················· | ···················· | ····················· | ···················· | ···················· | ···················· |
| J | 4451 | hCoV-19/England/MILK-9A56F3/2020\|EPI_ISL_550119\|20 | ···················· | ···················· | ····················· | ···················· | ···················· | ···················· |
| J | 4452 | hCoV-19/England/MILK-9A8B3D/2020\|EPI_ISL_550120\|20 | ···················· | ···················· | ····················· | ···················· | ···················· | ···················· |
| J | 4453 | hCoV-19/England/MILK-9A67D1/2020\|EPI_ISL_550122\|20 | ···················· | ···················· | ····················· | ···················· | ···················· | ···················· |
| J | 4454 | hCoV-19/England/MILK-9AA782/2020\|EPI_ISL_550800\|20 | ···················· | ···················· | ····················· | ···················· | ···················· | ···················· |
| J | 4455 | hCoV-19/England/MILK-9AA5F1/2020\|EPI_ISL_550801\|20 | ···················· | ···················· | ····················· | ···················· | ···················· | ···················· |
| J | 4456 | hCoV-19/England/MILK-9AA218/2020\|EPI_ISL_550802\|20 | ···················· | ···················· | ····················· | ···················· | ···················· | ···················· |
| J | 4457 | hCoV-19/England/MILK-9AA281/2020\|EPI_ISL_550803\|20 | ···················· | ···················· | ····················· | ···················· | ···················· | ···················· |
| J | 4458 | hCoV-19/England/MILK-9AA98C/2020\|EPI_ISL_550804\|20 | ···················· | ···················· | ····················· | ···················· | ···················· | ···················· |
| J | 4459 | hCoV-19/England/MILK-9AA245/2020\|EPI_ISL_550805\|20 | ···················· | ···················· | ····················· | ···················· | ······T············· | ···················· |
| J | 4460 | hCoV-19/England/MILK-9AA209/2020\|EPI_ISL_550806\|20 | ···················· | ···················· | ····················· | ···················· | ···················· | ···················· |
| J | 4461 | hCoV-19/England/MILK-9AA46D/2020\|EPI_ISL_550807\|20 | ···················· | ···················· | ····················· | ···················· | ···················· | ···················· |

| | | | | | | | | |
|---|---|---|---|---|---|---|---|---|
| J | 4462 | hCoV-19/England/MILK-9AA263/2020\|EPI_ISL_550808\|20 | ···················· | ···················· | ···················· | ···················· | ···················· | ···················· |
| J | 4463 | hCoV-19/England/MILK-9AA8F8/2020\|EPI_ISL_550809\|20 | ···················· | C··················· | ···················· | ···················· | ···················· | ···················· |
| J | 4464 | hCoV-19/England/MILK-9AA88F/2020\|EPI_ISL_550810\|20 | ···················· | ···················· | ···················· | ···················· | ···················· | ···················· |
| J | 4465 | hCoV-19/Wales/MILK-9AA6C1/2020\|EPI_ISL_550811\|2020 | ···················· | ···················· | ···················· | ···················· | ···················· | ···················· |
| J | 4466 | hCoV-19/England/MILK-9AACA1/2020\|EPI_ISL_550812\|20 | ···················· | ···················· | ···················· | ···················· | ···················· | ···················· |
| J | 4467 | hCoV-19/England/MILK-9AA0A5/2020\|EPI_ISL_550813\|20 | ···················· | ···················· | ···················· | ···················· | ···················· | ···················· |
| J | 4468 | hCoV-19/England/MILK-9A9FC7/2020\|EPI_ISL_550814\|20 | ···················· | ···················· | ···················· | ···················· | ···················· | ···················· |
| J | 4469 | hCoV-19/England/MILK-9AA904/2020\|EPI_ISL_550815\|20 | ···················· | ···················· | ···················· | ···················· | ···················· | ···················· |
| J | 4470 | hCoV-19/England/MILK-9AA56A/2020\|EPI_ISL_550816\|20 | ···················· | ···················· | ···················· | ···················· | ···················· | ···················· |
| J | 4471 | hCoV-19/England/MILK-9A9F9A/2020\|EPI_ISL_550817\|20 | ···················· | ···················· | ···················· | ···················· | ···················· | ···················· |
| J | 4472 | hCoV-19/England/MILK-9AACB0/2020\|EPI_ISL_550818\|20 | ···················· | ···················· | ···················· | ···················· | ···················· | ···················· |
| J | 4473 | hCoV-19/England/MILK-9AA3AC/2020\|EPI_ISL_550819\|20 | ···················· | ···················· | ···················· | ···················· | ···················· | ···················· |
| J | 4474 | hCoV-19/England/MILK-9A989F/2020\|EPI_ISL_550820\|20 | ···················· | ···················· | ···················· | ···················· | ···················· | ···················· |
| J | 4475 | hCoV-19/England/MILK-9AAA89/2020\|EPI_ISL_550821\|20 | ···················· | ···················· | ···················· | ···················· | ···················· | ···················· |
| J | 4476 | hCoV-19/England/MILK-9A9FE5/2020\|EPI_ISL_550822\|20 | ···················· | ···················· | ···················· | ···················· | ···················· | ···················· |
| J | 4477 | hCoV-19/England/MILK-9AA54C/2020\|EPI_ISL_550823\|20 | ···················· | ···················· | ···················· | ···················· | ···················· | ···················· |
| J | 4478 | hCoV-19/England/MILK-9AA148/2020\|EPI_ISL_550824\|20 | ···················· | ·A·················· | ···················· | ···················· | ···················· | ··A················· |
| J | 4479 | hCoV-19/Wales/MILK-9AA0E1/2020\|EPI_ISL_550825\|2020 | ···················· | ···················· | ···················· | ···················· | ···················· | ···················· |
| J | 4480 | hCoV-19/Wales/MILK-9AA658/2020\|EPI_ISL_550826\|2020 | ···················· | ···················· | ···················· | ···················· | ···················· | ···················· |
| J | 4481 | hCoV-19/England/MILK-9AA99B/2020\|EPI_ISL_550827\|20 | ···················· | ···················· | ···················· | ···················· | ···················· | ···················· |
| J | 4482 | hCoV-19/England/MILK-9AAC83/2020\|EPI_ISL_550828\|20 | ···················· | ···················· | ···················· | ···················· | ···················· | ···················· |
| J | 4483 | hCoV-19/England/MILK-9AA5C4/2020\|EPI_ISL_550829\|20 | ···················· | ···················· | ···················· | ···················· | ···················· | ···················· |
| J | 4484 | hCoV-19/England/MILK-9AAB0E/2020\|EPI_ISL_550830\|20 | ···················· | ···················· | ···················· | ···················· | ···················· | ···················· |
| J | 4485 | hCoV-19/England/MILK-9A9F8B/2020\|EPI_ISL_550831\|20 | ···················· | ···················· | ···················· | ···················· | ···················· | ···················· |
| J | 4486 | hCoV-19/England/MILK-9AA0D2/2020\|EPI_ISL_550832\|20 | ···················· | ···················· | ···················· | ···················· | ···················· | ···················· |
| J | 4487 | hCoV-19/England/MILK-9AAB2C/2020\|EPI_ISL_550833\|20 | ···················· | ···················· | ···················· | ···················· | ···················· | ···················· |
| J | 4488 | hCoV-19/Wales/MILK-9AA7EC/2020\|EPI_ISL_550834\|2020 | ···················· | ···················· | ···················· | ···················· | ···················· | ···················· |
| J | 4489 | hCoV-19/England/MILK-9AA087/2020\|EPI_ISL_550835\|20 | ···················· | ···················· | ···················· | ···················· | ···················· | ···················· |
| J | 4490 | hCoV-19/England/MILK-9AA193/2020\|EPI_ISL_550836\|20 | ···················· | ·····C·············· | ···················· | ···················· | ···················· | ···················· |
| J | 4491 | hCoV-19/England/MILK-9AA4C7/2020\|EPI_ISL_550837\|20 | ···················· | ···················· | ···················· | ···················· | ·····T·············· | ···················· |
| J | 4492 | hCoV-19/England/MILK-9AACED/2020\|EPI_ISL_550839\|20 | ···················· | ·A·················· | ···················· | ···················· | ···················· | ··A················· |
| J | 4493 | hCoV-19/England/MILK-9A9844/2020\|EPI_ISL_550841\|20 | ···················· | C··················· | ···················· | ···················· | ···················· | ···················· |
| J | 4494 | hCoV-19/England/MILK-9AA306/2020\|EPI_ISL_550842\|20 | ···················· | ·A·················· | ···················· | ···················· | ···················· | ··A················· |
| J | 4495 | hCoV-19/England/MILK-9AAB1D/2020\|EPI_ISL_550843\|20 | ···················· | ···················· | ···················· | ···················· | ···················· | ···················· |
| J | 4496 | hCoV-19/England/MILK-9AAB68/2020\|EPI_ISL_550844\|20 | ···················· | ···················· | ···················· | ···················· | ·····T·············· | ···················· |
| J | 4497 | hCoV-19/England/MILK-9AA51F/2020\|EPI_ISL_550845\|20 | ···················· | ···················· | ···················· | ···················· | ···················· | ···················· |
| J | 4498 | hCoV-19/Wales/MILK-9AA7DD/2020\|EPI_ISL_550846\|2020 | ···················· | ···················· | ···················· | ···················· | ···················· | ···················· |
| J | 4499 | hCoV-19/England/MILK-9AA0F0/2020\|EPI_ISL_550847\|20 | ···················· | ···················· | ···················· | ···················· | ···················· | ···················· |
| J | 4500 | hCoV-19/England/MILK-9AA12A/2020\|EPI_ISL_550848\|20 | ···················· | ···················· | ···················· | ···················· | ·····T·············· | ···················· |
| J | 4501 | hCoV-19/Wales/MILK-9AA1DF/2020\|EPI_ISL_550850\|2020 | ···················· | ···················· | ···················· | ···················· | ···················· | ···················· |
| J | 4502 | hCoV-19/England/MILK-9AA6A3/2020\|EPI_ISL_550851\|20 | ···················· | ···················· | ···················· | ···················· | ···················· | ···················· |
| J | 4503 | hCoV-19/England/MILK-9A9FF4/2020\|EPI_ISL_550852\|20 | ···················· | ···················· | ···················· | ···················· | ···················· | ···················· |
| J | 4504 | hCoV-19/England/MILK-9AA236/2020\|EPI_ISL_550853\|20 | ···················· | ···················· | ···················· | ···················· | ···················· | ···················· |
| J | 4505 | hCoV-19/England/MILK-9AAC92/2020\|EPI_ISL_550854\|20 | ···················· | ···················· | ···················· | ···················· | ···················· | ···················· |
| J | 4506 | hCoV-19/England/MILK-9AA2EB/2020\|EPI_ISL_550855\|20 | ···················· | ···················· | ···················· | ···················· | ···················· | ···················· |
| J | 4507 | hCoV-19/England/MILK-9A9871/2020\|EPI_ISL_550856\|20 | ···················· | ···················· | ···················· | ···················· | ···················· | ···················· |
| J | 4508 | hCoV-19/England/MILK-9AA7A0/2020\|EPI_ISL_550857\|20 | ···················· | ···················· | ···················· | ···················· | ···················· | ···················· |
| J | 4509 | hCoV-19/England/MILK-9AA53D/2020\|EPI_ISL_550858\|20 | ···················· | ···················· | ···················· | ···················· | ···················· | ···················· |
| J | 4510 | hCoV-19/England/MILK-9AA315/2020\|EPI_ISL_550859\|20 | ···················· | ·A·················· | ···················· | ···················· | ···················· | ··A················· |
| J | 4511 | hCoV-19/England/MILK-9AA157/2020\|EPI_ISL_550860\|20 | ···················· | ···················· | ···················· | ···················· | ···················· | ···················· |
| J | 4512 | hCoV-19/England/MILK-9AA421/2020\|EPI_ISL_550861\|20 | ···················· | ···················· | ···················· | ···················· | ···················· | ···················· |
| J | 4513 | hCoV-19/England/MILK-9AA48B/2020\|EPI_ISL_550862\|20 | ···················· | ···················· | ···················· | ···················· | ···················· | ···················· |
| J | 4514 | hCoV-19/England/MILK-9AAA3E/2020\|EPI_ISL_550863\|20 | ···················· | ···················· | ···················· | ···················· | ···················· | ···················· |
| J | 4515 | hCoV-19/England/MILK-9AA588/2020\|EPI_ISL_550864\|20 | ···················· | ···················· | ···················· | ···················· | ···················· | ···················· |
| J | 4516 | hCoV-19/England/MILK-9AADDB/2020\|EPI_ISL_550865\|20 | ···················· | ···················· | ···················· | ···················· | ···················· | ···················· |
| J | 4517 | hCoV-19/England/MILK-9A98CC/2020\|EPI_ISL_550866\|20 | ···················· | ···················· | ···················· | ···················· | ···················· | ···················· |
| J | 4518 | hCoV-19/England/MILK-9AA97D/2020\|EPI_ISL_550867\|20 | ···················· | ···················· | ···················· | ···················· | ···················· | ···················· |
| J | 4519 | hCoV-19/England/MILK-9AA500/2020\|EPI_ISL_550868\|20 | ···················· | ···················· | ···················· | ···················· | ···················· | ···················· |
| J | 4520 | hCoV-19/England/MILK-9AA1B1/2020\|EPI_ISL_550869\|20 | ···················· | ···················· | ···················· | ···················· | ·····T·············· | ···················· |
| J | 4521 | hCoV-19/England/MILK-9AA02D/2020\|EPI_ISL_550871\|20 | ···················· | ···················· | ···················· | ···················· | ···················· | ···················· |
| J | 4522 | hCoV-19/England/MILK-9A9FB8/2020\|EPI_ISL_550872\|20 | ···················· | ···················· | ···················· | ···················· | ···················· | ···················· |
| J | 4523 | hCoV-19/England/MILK-9A9FA9/2020\|EPI_ISL_550873\|20 | ···················· | ···················· | ···················· | ···················· | ···················· | ···················· |
| J | 4524 | hCoV-19/England/MILK-9AA825/2020\|EPI_ISL_550874\|20 | ···················· | ···················· | ···················· | ···················· | ···················· | ···················· |
| J | 4525 | hCoV-19/England/MILK-9AA166/2020\|EPI_ISL_550875\|20 | ···················· | ···················· | ···················· | ···················· | ·····T·············· | ···················· |
| J | 4526 | hCoV-19/England/MILK-9AAAC5/2020\|EPI_ISL_550876\|20 | ···················· | ···················· | ···················· | ···················· | ···················· | ···················· |
| J | 4527 | hCoV-19/England/MILK-9AA324/2020\|EPI_ISL_550877\|20 | ···················· | ···················· | ···················· | ···················· | ···················· | ···················· |
| J | 4528 | hCoV-19/England/MILK-9AAC1A/2020\|EPI_ISL_550878\|20 | ···················· | ···················· | ···················· | ···················· | ···················· | ···················· |
| J | 4529 | hCoV-19/England/MILK-9AA922/2020\|EPI_ISL_550879\|20 | ···················· | ···················· | ···················· | ···················· | ···················· | ···················· |
| J | 4530 | hCoV-19/England/MILK-9AA01E/2020\|EPI_ISL_550880\|20 | ···················· | ···················· | ···················· | ···················· | ···················· | ···················· |
| J | 4531 | hCoV-19/England/MILK-9AA89E/2020\|EPI_ISL_550881\|20 | ···················· | ···················· | ···················· | ···················· | ···················· | ···················· |
| J | 4532 | hCoV-19/England/ALDP-8FEDAC/2020\|EPI_ISL_551433\|20 | ···················· | ···················· | ···················· | ···················· | ···················· | ···················· |
| J | 4533 | hCoV-19/England/ALDP-916AD3/2020\|EPI_ISL_551435\|20 | ···················· | ·A·················· | ···················· | ···················· | ···················· | ···················· |
| J | 4534 | hCoV-19/England/ALDP-9364FD/2020\|EPI_ISL_551438\|20 | ···················· | ···················· | ···················· | ···················· | ···················· | ···················· |
| J | 4535 | hCoV-19/England/ALDP-92D392/2020\|EPI_ISL_551443\|20 | ···················· | ···················· | ···················· | ···················· | ···················· | ···················· |
| J | 4536 | hCoV-19/England/ALDP-923E8C/2020\|EPI_ISL_551444\|20 | ···················· | ···················· | ···················· | ···················· | ···················· | ···················· |
| J | 4537 | hCoV-19/England/ALDP-906186/2020\|EPI_ISL_551445\|20 | ···················· | ···················· | ···················· | ···················· | ···················· | ···················· |
| J | 4538 | hCoV-19/England/ALDP-905A6E/2020\|EPI_ISL_551446\|20 | ···················· | ···················· | ···················· | ···················· | ···················· | ···················· |
| J | 4539 | hCoV-19/England/ALDP-923FF2/2020\|EPI_ISL_551447\|20 | ···················· | ···················· | ···················· | ···················· | ···················· | ···················· |
| J | 4540 | hCoV-19/England/ALDP-90E607/2020\|EPI_ISL_551448\|20 | ···················· | ···················· | ···················· | ···················· | ···················· | ···················· |
| J | 4541 | hCoV-19/England/ALDP-9378E1/2020\|EPI_ISL_551449\|20 | ···················· | ···················· | ···················· | ···················· | ···················· | ···················· |

| | | | | | | | | |
|---|---|---|---|---|---|---|---|---|
| J | 4542 | hCoV-19/England/ALDP-93328D/2020\|EPI_ISL_551450\|20 | ···················· | ·A·················· | ···················· | ···················· | ···················· | ···················· |
| J | 4543 | hCoV-19/England/ALDP-9229AA/2020\|EPI_ISL_551451\|20 | ···················· | ···················· | ···················· | ···················· | ···················· | ···················· |
| J | 4544 | hCoV-19/England/ALDP-930A5B/2020\|EPI_ISL_551452\|20 | ···················· | ···················· | ···················· | ···················· | ···················· | ···················· |
| J | 4545 | hCoV-19/England/MILK-976752/2020\|EPI_ISL_552763\|20 | ···················· | ···················· | ···················· | ···················· | ···················· | ···················· |
| J | 4546 | hCoV-19/Wales/MILK-97689B/2020\|EPI_ISL_552765\|2020 | ···················· | ···················· | ···················· | ···················· | ···················· | ···················· |
| J | 4547 | hCoV-19/England/ALDP-981E40/2020\|EPI_ISL_552774\|20 | ···················· | ···················· | ···················· | ···················· | ···················· | ···················· |
| J | 4548 | hCoV-19/England/ALDP-981E9B/2020\|EPI_ISL_552776\|20 | ···················· | ···················· | ···················· | ···················· | ···················· | ···················· |
| J | 4549 | hCoV-19/England/ALDP-981D61/2020\|EPI_ISL_552779\|20 | ···················· | ···················· | ···················· | ···················· | ···················· | ···················· |
| J | 4550 | hCoV-19/England/ALDP-981C73/2020\|EPI_ISL_552780\|20 | ···················· | ···················· | ···················· | ···················· | ···················· | ···················· |
| J | 4551 | hCoV-19/England/MILK-97678F/2020\|EPI_ISL_552781\|20 | ···················· | ···················· | ···················· | ···················· | ···················· | ···················· |
| J | 4552 | hCoV-19/England/ALDP-981B94/2020\|EPI_ISL_552782\|20 | ···················· | ···················· | ···················· | ···················· | ···················· | ···················· |
| J | 4553 | hCoV-19/England/MILK-975E12/2020\|EPI_ISL_552784\|20 | ···················· | ···················· | ···················· | ···················· | ···················· | ···················· |
| J | 4554 | hCoV-19/England/ALDP-92D787/2020\|EPI_ISL_553000\|20 | ···················· | ···················· | ···················· | ···················· | ···················· | ···················· |
| J | 4555 | hCoV-19/England/ALDP-920D19/2020\|EPI_ISL_553001\|20 | ···················· | ···················· | ···················· | ···················· | ···················· | ···················· |
| J | 4556 | hCoV-19/England/ALDP-9358C5/2020\|EPI_ISL_553003\|20 | ···················· | ···················· | ···················· | ···················· | ···················· | ···················· |
| J | 4557 | hCoV-19/England/ALDP-9279E1/2020\|EPI_ISL_553004\|20 | ···················· | ···················· | ···················· | ···················· | ···················· | ···················· |
| J | 4558 | hCoV-19/England/MILK-97080A/2020\|EPI_ISL_553080\|20 | ···················· | ···················· | ···················· | ···················· | ···················· | ···················· |
| J | 4559 | hCoV-19/England/MILK-9709BC/2020\|EPI_ISL_553081\|20 | ···················· | ···················· | ···················· | ···················· | ···················· | ···················· |
| J | 4560 | hCoV-19/England/MILK-97098F/2020\|EPI_ISL_553083\|20 | ···················· | ····A··············· | ···················· | ···················· | ···················· | ···················· |
| J | 4561 | hCoV-19/England/MILK-9702C0/2020\|EPI_ISL_553086\|20 | ···················· | ···················· | ···················· | ···················· | ···················· | ···················· |
| J | 4562 | hCoV-19/Wales/MILK-9713EA/2020\|EPI_ISL_553087\|2020 | ···················· | ···················· | ···················· | ···················· | ···················· | ···················· |
| J | 4563 | hCoV-19/England/MILK-970934/2020\|EPI_ISL_553088\|20 | ···················· | ···················· | ···················· | ···················· | ···················· | ···················· |
| J | 4564 | hCoV-19/England/MILK-970CFF/2020\|EPI_ISL_553089\|20 | ···················· | ···················· | ···················· | ···················· | ···················· | ···················· |
| J | 4565 | hCoV-19/England/MILK-9710E3/2020\|EPI_ISL_553093\|20 | ···················· | ···················· | ···················· | ···················· | ···················· | ···················· |
| J | 4566 | hCoV-19/England/MILK-971238/2020\|EPI_ISL_553094\|20 | ···················· | ···················· | ···················· | ···················· | ···················· | ···················· |
| J | 4567 | hCoV-19/England/MILK-9703CD/2020\|EPI_ISL_553095\|20 | ···················· | ···················· | ···················· | ···················· | ···················· | ···················· |
| J | 4568 | hCoV-19/England/MILK-970B2F/2020\|EPI_ISL_553097\|20 | ········T··········· | ···················· | ···················· | ···················· | ···················· | ···················· |
| J | 4569 | hCoV-19/England/MILK-9718DC/2020\|EPI_ISL_553098\|20 | ···················· | ···················· | ···················· | ···················· | ···················· | ···················· |
| J | 4570 | hCoV-19/England/MILK-970D65/2020\|EPI_ISL_553099\|20 | ···················· | ···················· | ···················· | ···················· | ···················· | ···················· |
| J | 4571 | hCoV-19/England/ALDP-9556E3/2020\|EPI_ISL_554326\|20 | ···················· | ·A·················· | ···················· | ···················· | ···················· | ···················· |
| J | 4572 | hCoV-19/England/ALDP-95317B/2020\|EPI_ISL_554327\|20 | ···················· | ···················· | ···················· | ···················· | ···················· | ···················· |
| J | 4573 | hCoV-19/England/ALDP-950B61/2020\|EPI_ISL_554328\|20 | ···················· | ···················· | ···················· | ···················· | ···················· | ···················· |
| J | 4574 | hCoV-19/England/ALDP-953463/2020\|EPI_ISL_554329\|20 | ···················· | ···················· | ···················· | ···················· | ···················· | ···················· |
| J | 4575 | hCoV-19/England/ALDP-9509EF/2020\|EPI_ISL_554330\|20 | ···················· | ···················· | ···················· | ···················· | ···················· | ···················· |
| J | 4576 | hCoV-19/England/ALDP-9558ED/2020\|EPI_ISL_554332\|20 | ···················· | ···················· | ···················· | ···················· | ···················· | ···················· |
| J | 4577 | hCoV-19/England/ALDP-953D77/2020\|EPI_ISL_554334\|20 | ···················· | ···················· | ···················· | ···················· | ···················· | ···················· |
| J | 4578 | hCoV-19/England/ALDP-950D4D/2020\|EPI_ISL_554335\|20 | ···················· | ···················· | ···················· | ···················· | ···················· | ···················· |
| J | 4579 | hCoV-19/England/ALDP-94DB67/2020\|EPI_ISL_555656\|20 | ········T··········· | ···················· | ···················· | ···················· | ···················· | ···················· |
| J | 4580 | hCoV-19/England/ALDP-94DE5F/2020\|EPI_ISL_555657\|20 | ···················· | ···················· | ···················· | ···················· | ···················· | ···················· |
| J | 4581 | hCoV-19/England/ALDP-9523A3/2020\|EPI_ISL_555658\|20 | ···················· | ···················· | ···················· | ···················· | ···················· | ···················· |
| J | 4582 | hCoV-19/England/ALDP-94DBB2/2020\|EPI_ISL_555659\|20 | ···················· | ···················· | ···················· | ···················· | ···················· | ···················· |
| J | 4583 | hCoV-19/England/ALDP-94480F/2020\|EPI_ISL_555662\|20 | ···················· | ···················· | ···················· | ···················· | ···················· | ···················· |
| J | 4584 | hCoV-19/England/ALDP-94E3E6/2020\|EPI_ISL_555664\|20 | ···················· | ···················· | ···················· | ···················· | ···················· | ···················· |
| J | 4585 | hCoV-19/England/ALDP-95AB94/2020\|EPI_ISL_555666\|20 | ···················· | ···················· | ···················· | ···················· | ···················· | ···················· |
| J | 4586 | hCoV-19/England/ALDP-8D19D8/2020\|EPI_ISL_556987\|20 | ···················· | ···················· | ···················· | ···················· | ···················· | ···················· |
| J | 4587 | hCoV-19/England/ALDP-8FC852/2020\|EPI_ISL_556988\|20 | ···················· | ···················· | ···················· | ···················· | ···················· | ···················· |
| J | 4588 | hCoV-19/England/ALDP-8D85EC/2020\|EPI_ISL_556989\|20 | ···················· | ···················· | ···················· | ···················· | ···················· | ···················· |
| J | 4589 | hCoV-19/England/ALDP-8DF7D0/2020\|EPI_ISL_556991\|20 | ···················· | ···················· | ···················· | ···················· | ···················· | ···················· |
| J | 4590 | hCoV-19/England/ALDP-8E036F/2020\|EPI_ISL_556992\|20 | ···················· | ···················· | ···················· | ···················· | ···················· | ···················· |
| J | 4591 | hCoV-19/England/ALDP-8FEF00/2020\|EPI_ISL_556993\|20 | ···················· | ···················· | ···················· | ···················· | ···················· | ···················· |
| J | 4592 | hCoV-19/England/ALDP-8DEDFD/2020\|EPI_ISL_556994\|20 | ···················· | ···················· | ···················· | ···················· | ···················· | ···················· |
| J | 4593 | hCoV-19/England/ALDP-8D8379/2020\|EPI_ISL_556995\|20 | ···················· | ···················· | ···················· | ···················· | ···················· | ···················· |
| J | 4594 | hCoV-19/England/ALDP-8DF4E8/2020\|EPI_ISL_556996\|20 | ···················· | ···················· | ···················· | ···················· | ···················· | ···················· |
| J | 4595 | hCoV-19/England/ALDP-8FF163/2020\|EPI_ISL_556997\|20 | ···················· | ·A·················· | ···················· | ···················· | ···················· | ···················· |
| J | 4596 | hCoV-19/England/ALDP-5092FD/2020\|EPI_ISL_559200\|20 | ···················· | ···················· | ···················· | ···················· | ···················· | ···················· |
| J | 4597 | hCoV-19/England/MILK-4B3378/2020\|EPI_ISL_559201\|20 | ···················· | ···················· | ···················· | ···················· | ···················· | ···················· |
| J | 4598 | hCoV-19/England/ALDP-4F4F3B/2020\|EPI_ISL_559205\|20 | ···················· | ···················· | ···················· | ···················· | ···················· | ···················· |
| J | 4599 | hCoV-19/England/ALDP-5026F8/2020\|EPI_ISL_559208\|20 | ···················· | ···················· | ···················· | ···················· | ···················· | ···················· |
| J | 4600 | hCoV-19/England/ALDP-4FA375/2020\|EPI_ISL_559209\|20 | ···················· | ···················· | ···················· | ···················· | ···················· | ···················· |
| J | 4601 | hCoV-19/England/ALDP-508DC0/2020\|EPI_ISL_559210\|20 | ···················· | ···················· | ···················· | ···················· | ···················· | ···················· |
| J | 4602 | hCoV-19/England/ALDP-4FC1C4/2020\|EPI_ISL_559211\|20 | ···················· | ···················· | ···················· | ···················· | ···················· | ···················· |
| J | 4603 | hCoV-19/England/ALDP-4F48F4/2020\|EPI_ISL_559212\|20 | ···················· | ···················· | ···················· | ···················· | ···················· | ···················· |
| J | 4604 | hCoV-19/England/ALDP-500144/2020\|EPI_ISL_559214\|20 | ···················· | ···················· | ···················· | ···················· | ···················· | ···················· |
| J | 4605 | hCoV-19/England/ALDP-4F7F0B/2020\|EPI_ISL_559216\|20 | ···················· | ···················· | ···················· | ···················· | ···················· | ···················· |
| J | 4606 | hCoV-19/England/ALDP-4F6A56/2020\|EPI_ISL_559217\|20 | ···················· | ···················· | ···················· | ···················· | ···················· | ···················· |
| J | 4607 | hCoV-19/England/MILK-4C414D/2020\|EPI_ISL_559218\|20 | ···················· | ···················· | ···················· | ···················· | ···················· | ···················· |
| J | 4608 | hCoV-19/England/ALDP-4F7105/2020\|EPI_ISL_559219\|20 | ···················· | ···················· | ···················· | ···················· | ···················· | ···················· |
| J | 4609 | hCoV-19/England/QEUH-9B8499/2020\|EPI_ISL_549800\|20 | ···················· | ···················· | ···················· | ···················· | ···················· | ···················· |
| J | 4610 | hCoV-19/England/QEUH-9B9EC7/2020\|EPI_ISL_549801\|20 | ···················· | ···················· | ···················· | ···················· | ······T············· | ···················· |
| J | 4611 | hCoV-19/England/QEUH-9B8FC5/2020\|EPI_ISL_549802\|20 | ···················· | ···················· | ···················· | ···················· | ···················· | ···················· |
| J | 4612 | hCoV-19/England/QEUH-9B8CCE/2020\|EPI_ISL_549803\|20 | ···················· | ···················· | ···················· | ···················· | ···················· | ···················· |
| J | 4613 | hCoV-19/England/QEUH-9B8E04/2020\|EPI_ISL_549805\|20 | ···················· | ···················· | ···················· | ···················· | ···················· | ···················· |
| J | 4614 | hCoV-19/England/QEUH-9B929E/2020\|EPI_ISL_549806\|20 | ···················· | ···················· | ···················· | ···················· | ···················· | ···················· |
| J | 4615 | hCoV-19/England/MILK-99D45E/2020\|EPI_ISL_549808\|20 | ···················· | ···················· | ···················· | ···················· | ···················· | ···················· |
| J | 4616 | hCoV-19/England/MILK-99CE06/2020\|EPI_ISL_549809\|20 | ···················· | ···················· | ···················· | ···················· | ···················· | ···················· |
| J | 4617 | hCoV-19/England/MILK-99CF5E/2020\|EPI_ISL_549810\|20 | ···················· | ···················· | ···················· | ···················· | ···················· | ···················· |
| J | 4618 | hCoV-19/England/MILK-99CF6D/2020\|EPI_ISL_549811\|20 | ···················· | ···················· | ···················· | ···················· | ···················· | ···················· |
| J | 4619 | hCoV-19/England/MILK-99CFD6/2020\|EPI_ISL_549812\|20 | ···················· | ···················· | ···················· | ···················· | ···················· | ···················· |
| J | 4620 | hCoV-19/England/MILK-99D6FE/2020\|EPI_ISL_549813\|20 | ···················· | ·A·················· | ···················· | ···················· | ···················· | ···················· |
| J | 4621 | hCoV-19/England/MILK-99D139/2020\|EPI_ISL_549814\|20 | ···················· | ···················· | ···················· | ···················· | ···················· | ···················· |

| | | | | | | | |
|---|---|---|---|---|---|---|---|
| J | 4622 hCoV-19/England/ALDP-935E4B/2020\|EPI_ISL_553005\|20 | ···················· | ···················· | ····················· | ···················· | ···················· | ···················· |
| J | 4623 hCoV-19/England/ALDP-913809/2020\|EPI_ISL_553006\|20 | ···················· | ···················· | ····················· | ···················· | ···················· | ···················· |
| J | 4624 hCoV-19/Wales/MILK-97172A/2020\|EPI_ISL_553008\|2020 | ···················· | ···················· | ····················· | ···················· | ···················· | ···················· |
| J | 4625 hCoV-19/England/MILK-9704BB/2020\|EPI_ISL_553009\|20 | ···················· | ···················· | ····················· | ···················· | ···················· | ···················· |
| J | 4626 hCoV-19/England/MILK-970B4D/2020\|EPI_ISL_553010\|20 | ···················· | ···················· | ····················· | ···················· | ···················· | ···················· |
| J | 4627 hCoV-19/England/MILK-970B10/2020\|EPI_ISL_553011\|20 | ···················· | ···················· | ····················· | ···················· | ···················· | ···················· |
| J | 4628 hCoV-19/England/MILK-970E62/2020\|EPI_ISL_553012\|20 | ···················· | ···················· | ····················· | ···················· | ···················· | ···················· |
| J | 4629 hCoV-19/England/MILK-970A6E/2020\|EPI_ISL_553014\|20 | ···················· | ···················· | ····················· | ···················· | ···················· | ···················· |
| J | 4630 hCoV-19/Wales/MILK-9713DB/2020\|EPI_ISL_553015\|2020 | ···················· | ···················· | ····················· | ···················· | ···················· | ···················· |
| J | 4631 hCoV-19/Wales/MILK-970B98/2020\|EPI_ISL_553016\|2020 | ···················· | ···················· | ····················· | ···················· | ···················· | ···················· |
| J | 4632 hCoV-19/England/MILK-970433/2020\|EPI_ISL_553017\|20 | ···················· | ···················· | ····················· | ···················· | ···················· | ···················· |
| J | 4633 hCoV-19/England/MILK-971678/2020\|EPI_ISL_553018\|20 | ···················· | ···················· | ····················· | ···················· | ···················· | ···················· |
| J | 4634 hCoV-19/England/MILK-9714F6/2020\|EPI_ISL_553019\|20 | ···················· | ···················· | ····················· | ···················· | ···················· | ···················· |
| J | 4635 hCoV-19/England/MILK-9703BE/2020\|EPI_ISL_553020\|20 | ···················· | ·A·················· | ····················· | ···················· | ···················· | ··A················· |
| J | 4636 hCoV-19/England/MILK-9714BA/2020\|EPI_ISL_553021\|20 | ···················· | ···················· | ····················· | ···················· | ···················· | ···················· |
| J | 4637 hCoV-19/Wales/MILK-970A5F/2020\|EPI_ISL_553023\|2020 | ···················· | ···················· | ····················· | ···················· | ···················· | ···················· |
| J | 4638 hCoV-19/England/MILK-970DA1/2020\|EPI_ISL_553024\|20 | ···················· | ···················· | ····················· | ···················· | ···················· | ···················· |
| J | 4639 hCoV-19/England/MILK-971915/2020\|EPI_ISL_553026\|20 | ···················· | ···················· | ····················· | ···················· | ···················· | ···················· |
| J | 4640 hCoV-19/England/MILK-9702A2/2020\|EPI_ISL_553027\|20 | ···················· | ···················· | ····················· | ···················· | ···················· | ···················· |
| J | 4641 hCoV-19/England/MILK-9711A4/2020\|EPI_ISL_553028\|20 | ···················· | ···················· | ····················· | ···················· | ···················· | ···················· |
| J | 4642 hCoV-19/Wales/MILK-9712B0/2020\|EPI_ISL_553029\|2020 | ···················· | ···················· | ····················· | ···················· | ···················· | ···················· |
| J | 4643 hCoV-19/England/MILK-9718BE/2020\|EPI_ISL_553031\|20 | ···················· | ···················· | ····················· | ···················· | ···················· | ···················· |
| J | 4644 hCoV-19/England/MILK-971836/2020\|EPI_ISL_553032\|20 | ···················· | ···················· | ····················· | ···················· | ···················· | ···················· |
| J | 4645 hCoV-19/England/MILK-970C68/2020\|EPI_ISL_553033\|20 | ···················· | ···················· | ····················· | ···················· | ···················· | ···················· |
| J | 4646 hCoV-19/England/MILK-97071C/2020\|EPI_ISL_553034\|20 | ···················· | ···················· | ····················· | ···················· | ···················· | ···················· |
| J | 4647 hCoV-19/England/MILK-970A9B/2020\|EPI_ISL_553035\|20 | ···················· | ···················· | ····················· | ···················· | ···················· | ···················· |
| J | 4648 hCoV-19/England/MILK-970D38/2020\|EPI_ISL_553036\|20 | ···················· | ···················· | ····················· | ···················· | ···················· | ···················· |
| J | 4649 hCoV-19/England/MILK-970C2C/2020\|EPI_ISL_553040\|20 | ···················· | ···················· | ····················· | ···················· | ···················· | ···················· |
| J | 4650 hCoV-19/England/MILK-970E71/2020\|EPI_ISL_553041\|20 | ···················· | ···················· | ···········T········· | ···················· | ···················· | ···················· |
| J | 4651 hCoV-19/England/MILK-9702DF/2020\|EPI_ISL_553042\|20 | ···················· | ···················· | ····················· | ···················· | ···················· | ···················· |
| J | 4652 hCoV-19/England/MILK-9710D4/2020\|EPI_ISL_553043\|20 | ···················· | ···················· | ····················· | ···················· | ···················· | ···················· |
| J | 4653 hCoV-19/England/MILK-97099E/2020\|EPI_ISL_553046\|20 | ···················· | ···A················ | ····················· | ···················· | ···················· | ···················· |
| J | 4654 hCoV-19/England/MILK-970EF9/2020\|EPI_ISL_553047\|20 | ···················· | ···················· | ····················· | ···················· | ···················· | ···················· |
| J | 4655 hCoV-19/England/MILK-970846/2020\|EPI_ISL_553048\|20 | ···················· | ···················· | ····················· | ···················· | ···················· | ···················· |
| J | 4656 hCoV-19/England/MILK-970D1A/2020\|EPI_ISL_553049\|20 | ···················· | ···················· | ····················· | ···················· | ···················· | ···················· |
| J | 4657 hCoV-19/England/MILK-97106B/2020\|EPI_ISL_553051\|20 | ···················· | ···················· | ····················· | ···················· | ···················· | ···················· |
| J | 4658 hCoV-19/England/MILK-9711C2/2020\|EPI_ISL_553052\|20 | ···················· | ···················· | ····················· | ···················· | ···················· | ···················· |
| J | 4659 hCoV-19/England/MILK-9716E1/2020\|EPI_ISL_553053\|20 | ···················· | ···················· | ····················· | ···················· | ···················· | ···················· |
| J | 4660 hCoV-19/England/MILK-970C95/2020\|EPI_ISL_553056\|20 | ···················· | ·A·················· | ····················· | ···················· | ···················· | ···················· |
| J | 4661 hCoV-19/England/MILK-970C3B/2020\|EPI_ISL_553057\|20 | ···················· | ···················· | ····················· | ···················· | ···················· | ···················· |
| J | 4662 hCoV-19/England/MILK-9704E8/2020\|EPI_ISL_553059\|20 | ···················· | ···················· | ····················· | ···················· | ···················· | ···················· |
| J | 4663 hCoV-19/England/MILK-970C0E/2020\|EPI_ISL_553061\|20 | ···················· | ···················· | ····················· | ···················· | ···················· | ···················· |
| J | 4664 hCoV-19/England/MILK-970819/2020\|EPI_ISL_553062\|20 | ···················· | ···················· | ····················· | ···················· | ···················· | ···················· |
| J | 4665 hCoV-19/England/MILK-97170C/2020\|EPI_ISL_553063\|20 | ···················· | ···················· | ····················· | ···················· | ···················· | ···················· |
| J | 4666 hCoV-19/England/MILK-97165A/2020\|EPI_ISL_553064\|20 | ···················· | ···················· | ····················· | ···················· | ···················· | ···················· |
| J | 4667 hCoV-19/England/MILK-97048E/2020\|EPI_ISL_553065\|20 | ···················· | ···················· | ····················· | ···················· | ···················· | ···················· |
| J | 4668 hCoV-19/England/MILK-97021B/2020\|EPI_ISL_553066\|20 | ···················· | ···················· | ····················· | ···················· | ···················· | ···················· |
| J | 4669 hCoV-19/England/MILK-97107A/2020\|EPI_ISL_553067\|20 | ···················· | ·A·················· | ····················· | ···················· | ···················· | ···················· |
| J | 4670 hCoV-19/England/MILK-970363/2020\|EPI_ISL_553072\|20 | ···················· | ···················· | ····················· | ···················· | ···················· | ···················· |
| J | 4671 hCoV-19/England/MILK-970390/2020\|EPI_ISL_553073\|20 | ···················· | ···················· | ····················· | ···················· | ···················· | ···················· |
| J | 4672 hCoV-19/England/MILK-97149C/2020\|EPI_ISL_553074\|20 | ···················· | ···················· | ····················· | ···················· | ···················· | ···················· |
| J | 4673 hCoV-19/England/MILK-971098/2020\|EPI_ISL_553075\|20 | ···T················ | ···················· | ····················· | ···················· | ···················· | ···················· |
| J | 4674 hCoV-19/England/MILK-971775/2020\|EPI_ISL_553076\|20 | ···················· | ···················· | ····················· | ···················· | ···················· | ···················· |
| J | 4675 hCoV-19/England/MILK-97065B/2020\|EPI_ISL_553077\|20 | ···················· | ···················· | ····················· | ···················· | ···················· | ···················· |
| J | 4676 hCoV-19/England/MILK-970EBD/2020\|EPI_ISL_553079\|20 | ···················· | ···················· | ····················· | ···················· | ···················· | ···················· |
| J | 4677 hCoV-19/England/MILK-970952/2020\|EPI_ISL_553101\|20 | ···················· | ···A················ | ····················· | ···················· | ···················· | ···················· |
| J | 4678 hCoV-19/England/MILK-97057C/2020\|EPI_ISL_553102\|20 | ···················· | ···················· | ····················· | ···················· | ···················· | ···················· |
| J | 4679 hCoV-19/Wales/MILK-970FF6/2020\|EPI_ISL_553103\|2020 | ···················· | ···················· | ····················· | ···················· | ···················· | ···················· |
| J | 4680 hCoV-19/England/MILK-971326/2020\|EPI_ISL_553106\|20 | ···················· | ···················· | ····················· | ···················· | ···················· | ···················· |
| J | 4681 hCoV-19/England/MILK-97158A/2020\|EPI_ISL_553107\|20 | ···················· | ···················· | ····················· | ···················· | ···················· | ···················· |
| J | 4682 hCoV-19/England/MILK-9711E0/2020\|EPI_ISL_553111\|20 | ···················· | ···················· | ····················· | ···················· | ···················· | ···················· |
| J | 4683 hCoV-19/England/MILK-9701D2/2020\|EPI_ISL_553112\|20 | ···················· | ···················· | ····················· | ···················· | ···················· | ···················· |
| J | 4684 hCoV-19/Wales/MILK-971195/2020\|EPI_ISL_553113\|2020 | ···················· | ···················· | ····················· | ···················· | ···················· | ···················· |
| J | 4685 hCoV-19/England/MILK-970E44/2020\|EPI_ISL_553114\|20 | ···················· | ···················· | ····················· | ···················· | ···················· | ···················· |
| J | 4686 hCoV-19/England/ALDP-9511E6/2020\|EPI_ISL_554336\|20 | ···················· | ···················· | ····················· | ···················· | ···················· | ···················· |
| J | 4687 hCoV-19/England/ALDP-9557B3/2020\|EPI_ISL_554339\|20 | ···················· | ···················· | ····················· | ···················· | ···················· | ···················· |
| J | 4688 hCoV-19/England/ALDP-9532A5/2020\|EPI_ISL_554343\|20 | ···················· | ···················· | ····················· | ···················· | ···················· | ···················· |
| J | 4689 hCoV-19/England/ALDP-950ABF/2020\|EPI_ISL_554344\|20 | ···················· | ···················· | ····················· | ···················· | ···················· | ···················· |
| J | 4690 hCoV-19/England/ALDP-95367C/2020\|EPI_ISL_554345\|20 | ···················· | ···················· | ····················· | ···················· | ···················· | ···················· |
| J | 4691 hCoV-19/England/ALDP-954004/2020\|EPI_ISL_554347\|20 | ···················· | ···················· | ····················· | ···················· | ···················· | ···················· |
| J | 4692 hCoV-19/England/ALDP-955865/2020\|EPI_ISL_554348\|20 | ···················· | ···················· | ····················· | ···················· | ···················· | ···················· |
| J | 4693 hCoV-19/England/ALDP-951289/2020\|EPI_ISL_554349\|20 | ···················· | ···················· | ····················· | ···················· | ···················· | ···················· |
| J | 4694 hCoV-19/England/ALDP-95565C/2020\|EPI_ISL_554351\|20 | ···················· | ···················· | ····················· | ···················· | ···················· | ···················· |
| J | 4695 hCoV-19/England/ALDP-95332A/2020\|EPI_ISL_554352\|20 | ···················· | ···················· | ····················· | ···················· | ···················· | ···················· |
| J | 4696 hCoV-19/England/ALDP-9512A7/2020\|EPI_ISL_554354\|20 | ···················· | ···················· | ····················· | ···················· | ···················· | ···················· |
| J | 4697 hCoV-19/England/ALDP-95375B/2020\|EPI_ISL_554355\|20 | ···················· | ···················· | ····················· | ···················· | ···················· | ···················· |
| J | 4698 hCoV-19/England/ALDP-9511C8/2020\|EPI_ISL_554356\|20 | ···················· | ···················· | ····················· | ···················· | ···················· | ···················· |
| J | 4699 hCoV-19/England/ALDP-953DA4/2020\|EPI_ISL_554357\|20 | ···················· | ···················· | ····················· | ···················· | ···················· | ···················· |
| J | 4700 hCoV-19/England/ALDP-950C8C/2020\|EPI_ISL_554360\|20 | ···················· | ···················· | ····················· | ···················· | ···················· | ···················· |
| J | 4701 hCoV-19/England/ALDP-950A73/2020\|EPI_ISL_554361\|20 | ···················· | ···················· | ····················· | ···················· | ···················· | ···················· |

| | | | | | | | | |
|---|---|---|---|---|---|---|---|---|
| J | 4702 | hCoV-19/England/ALDP-9511D7/2020\|EPI_ISL_554362\|20 | ···················· | ···················· | ····················· | ···················· | ···················· | ···················· |
| J | 4703 | hCoV-19/England/ALDP-950B8F/2020\|EPI_ISL_554363\|20 | ···················· | ···················· | ····················· | ···················· | ···················· | ···················· |
| J | 4704 | hCoV-19/England/ALDP-953418/2020\|EPI_ISL_554364\|20 | ···················· | ···················· | ····················· | ···················· | ···················· | ···················· |
| J | 4705 | hCoV-19/England/ALDP-950BF8/2020\|EPI_ISL_554365\|20 | ···················· | ···················· | ····················· | ···················· | ···················· | ···················· |
| J | 4706 | hCoV-19/England/ALDP-95374C/2020\|EPI_ISL_554368\|20 | ···················· | ···················· | ····················· | ···················· | ···················· | ···················· |
| J | 4707 | hCoV-19/England/ALDP-955908/2020\|EPI_ISL_554369\|20 | ···················· | ···················· | ····················· | ···················· | ···················· | ···················· |
| J | 4708 | hCoV-19/England/ALDP-950D5C/2020\|EPI_ISL_554370\|20 | ···················· | ···················· | ····················· | ···················· | ···················· | ···················· |
| J | 4709 | hCoV-19/England/ALDP-9537D3/2020\|EPI_ISL_554371\|20 | ···················· | ···················· | ····················· | ···················· | ···················· | ···················· |
| J | 4710 | hCoV-19/England/ALDP-9556A7/2020\|EPI_ISL_554372\|20 | ···················· | ···················· | ····················· | ···················· | ···················· | ···················· |
| J | 4711 | hCoV-19/England/ALDP-955768/2020\|EPI_ISL_554373\|20 | ···················· | ···················· | ····················· | ···················· | ···················· | ···················· |
| J | 4712 | hCoV-19/England/ALDP-954374/2020\|EPI_ISL_554374\|20 | ···················· | ···················· | ····················· | ···················· | ···················· | ···················· |
| J | 4713 | hCoV-19/England/ALDP-9537E2/2020\|EPI_ISL_554376\|20 | ···················· | ···················· | ····················· | ···················· | ···················· | ···················· |
| J | 4714 | hCoV-19/England/ALDP-955838/2020\|EPI_ISL_554377\|20 | ···················· | ···················· | ····················· | ···················· | ···················· | ···················· |
| J | 4715 | hCoV-19/England/ALDP-950800/2020\|EPI_ISL_554379\|20 | ···················· | ···················· | ····················· | ···················· | ···················· | ···················· |
| J | 4716 | hCoV-19/England/ALDP-9556D4/2020\|EPI_ISL_554380\|20 | ···················· | ···················· | ····················· | ···················· | ···················· | ···················· |
| J | 4717 | hCoV-19/England/ALDP-955883/2020\|EPI_ISL_554381\|20 | ···················· | ···················· | ····················· | ···················· | ···················· | ···················· |
| J | 4718 | hCoV-19/England/ALDP-950A46/2020\|EPI_ISL_554383\|20 | ···················· | ···················· | ····················· | ···················· | ···················· | ···················· |
| J | 4719 | hCoV-19/England/ALDP-9514FC/2020\|EPI_ISL_554384\|20 | ···················· | ···················· | ····················· | ···················· | ···················· | ···················· |
| J | 4720 | hCoV-19/Wales/ALDP-95572C/2020\|EPI_ISL_554387\|2020 | ···················· | ···················· | ····················· | ···················· | ···················· | ···················· |
| J | 4721 | hCoV-19/England/ALDP-95323C/2020\|EPI_ISL_554388\|20 | ···················· | ···················· | ····················· | ···················· | ···················· | ···················· |
| J | 4722 | hCoV-19/England/ALDP-950730/2020\|EPI_ISL_554389\|20 | ···················· | ···················· | ····················· | ···················· | ···················· | ···················· |
| J | 4723 | hCoV-19/England/ALDP-95167E/2020\|EPI_ISL_554391\|20 | ···················· | ···················· | ····················· | ···················· | ···················· | ···················· |
| J | 4724 | hCoV-19/England/ALDP-955698/2020\|EPI_ISL_554392\|20 | ···················· | ···················· | ····················· | ···················· | ···················· | ···················· |
| J | 4725 | hCoV-19/England/ALDP-950976/2020\|EPI_ISL_554393\|20 | ···················· | ···················· | ····················· | ···················· | ···················· | ···················· |
| J | 4726 | hCoV-19/England/ALDP-953E47/2020\|EPI_ISL_554394\|20 | ···················· | ···················· | ····················· | ···················· | ···················· | ···················· |
| J | 4727 | hCoV-19/England/ALDP-9507A9/2020\|EPI_ISL_554395\|20 | ···················· | ···················· | ····················· | ···················· | ···················· | ···················· |
| J | 4728 | hCoV-19/England/ALDP-950C5F/2020\|EPI_ISL_554396\|20 | ···················· | ···················· | ····················· | ···················· | ···················· | ···················· |
| J | 4729 | hCoV-19/England/ALDP-9508E2/2020\|EPI_ISL_554397\|20 | ···················· | ·A·················· | ····················· | ···················· | ···················· | ···················· |
| J | 4730 | hCoV-19/England/ALDP-953278/2020\|EPI_ISL_554398\|20 | ···················· | ···················· | ····················· | ···················· | ···················· | ···················· |
| J | 4731 | hCoV-19/England/ALDP-953779/2020\|EPI_ISL_554399\|20 | ···················· | ···················· | ····················· | ···················· | ···················· | ···················· |
| J | 4732 | hCoV-19/England/ALDP-95368B/2020\|EPI_ISL_554401\|20 | ···················· | ·A·················· | ····················· | ···················· | ···················· | ···················· |
| J | 4733 | hCoV-19/England/ALDP-953393/2020\|EPI_ISL_554402\|20 | ···················· | ···················· | ····················· | ···················· | ···················· | ···················· |
| J | 4734 | hCoV-19/England/ALDP-955874/2020\|EPI_ISL_554404\|20 | ···················· | ···················· | ····················· | ···················· | ···················· | ···················· |
| J | 4735 | hCoV-19/England/ALDP-950C7D/2020\|EPI_ISL_554405\|20 | ···················· | ···················· | ····················· | ···················· | ···················· | ···················· |
| J | 4736 | hCoV-19/England/ALDP-95127A/2020\|EPI_ISL_554406\|20 | ···················· | ···················· | ····················· | ···················· | ···················· | ···············A···· |
| J | 4737 | hCoV-19/England/ALDP-9536B8/2020\|EPI_ISL_554407\|20 | ···················· | ···················· | ····················· | ···················· | ···················· | ···················· |
| J | 4738 | hCoV-19/England/ALDP-952F54/2020\|EPI_ISL_554408\|20 | ···················· | ···················· | ····················· | ···················· | ···················· | ···················· |
| J | 4739 | hCoV-19/England/ALDP-953982/2020\|EPI_ISL_554410\|20 | ···················· | ···················· | ····················· | ···················· | ···················· | ···················· |
| J | 4740 | hCoV-19/England/ALDP-953DE0/2020\|EPI_ISL_554411\|20 | ···················· | ···················· | ····················· | ···················· | ···················· | ···················· |
| J | 4741 | hCoV-19/England/ALDP-953357/2020\|EPI_ISL_554412\|20 | ···················· | ···················· | ····················· | ···················· | ···················· | ···················· |
| J | 4742 | hCoV-19/England/ALDP-95316C/2020\|EPI_ISL_554413\|20 | ···················· | ···················· | ····················· | ···················· | ···················· | ···················· |
| J | 4743 | hCoV-19/England/ALDP-954268/2020\|EPI_ISL_554414\|20 | ···················· | ···················· | ····················· | ···················· | ···················· | ···················· |
| J | 4744 | hCoV-19/England/ALDP-9509C1/2020\|EPI_ISL_554415\|20 | ···················· | ···················· | ····················· | ···················· | ···················· | ···················· |
| J | 4745 | hCoV-19/England/ALDP-95366D/2020\|EPI_ISL_554416\|20 | ···················· | ···················· | ····················· | ···················· | ···················· | ···················· |
| J | 4746 | hCoV-19/England/ALDP-955452/2020\|EPI_ISL_554417\|20 | ···················· | ···················· | ····················· | ···················· | ···················· | ···················· |
| J | 4747 | hCoV-19/England/ALDP-95141A/2020\|EPI_ISL_554418\|20 | ···················· | ···················· | ····················· | ···················· | ···················· | ···················· |
| J | 4748 | hCoV-19/England/ALDP-950B70/2020\|EPI_ISL_554419\|20 | ···················· | ···················· | ····················· | ···················· | ···················· | ···················· |
| J | 4749 | hCoV-19/England/ALDP-9537F1/2020\|EPI_ISL_554421\|20 | ···················· | ···················· | ····················· | ···················· | ···················· | ···················· |
| J | 4750 | hCoV-19/England/ALDP-9508A6/2020\|EPI_ISL_554422\|20 | ···················· | ···················· | ····················· | ···················· | ···················· | ···················· |
| J | 4751 | hCoV-19/England/ALDP-9514ED/2020\|EPI_ISL_554423\|20 | ···················· | ···················· | ····················· | ···················· | ···················· | ···················· |
| J | 4752 | hCoV-19/England/ALDP-950AEC/2020\|EPI_ISL_554424\|20 | ···················· | ···················· | ····················· | ···················· | ···················· | ···················· |
| J | 4753 | hCoV-19/England/ALDP-9558A1/2020\|EPI_ISL_554425\|20 | ···················· | ···················· | ····················· | ···················· | ···················· | ···················· |
| J | 4754 | hCoV-19/England/ALDP-951447/2020\|EPI_ISL_554426\|20 | ···················· | ···················· | ····················· | ···················· | ···················· | ···················· |
| J | 4755 | hCoV-19/England/ALDP-9534BE/2020\|EPI_ISL_554427\|20 | ···················· | ···················· | ····················· | ···················· | ···················· | ···················· |
| J | 4756 | hCoV-19/England/ALDP-950C04/2020\|EPI_ISL_554428\|20 | ···················· | ···················· | ····················· | ···················· | ···················· | ···················· |
| J | 4757 | hCoV-19/England/ALDP-9557A4/2020\|EPI_ISL_554429\|20 | ···················· | ···················· | ····················· | ···················· | ···················· | ···················· |
| J | 4758 | hCoV-19/England/ALDP-953CE3/2020\|EPI_ISL_554430\|20 | ···················· | ···················· | ····················· | ···················· | ···················· | ···················· |
| J | 4759 | hCoV-19/England/ALDP-953E0B/2020\|EPI_ISL_554431\|20 | ···················· | ···················· | ····················· | ···················· | ···················· | ···················· |
| J | 4760 | hCoV-19/England/ALDP-950985/2020\|EPI_ISL_554432\|20 | ···················· | ···················· | ····················· | ···················· | ···················· | ···················· |
| J | 4761 | hCoV-19/England/ALDP-9507F4/2020\|EPI_ISL_554434\|20 | ···················· | ···················· | ····················· | ···················· | ···················· | ···················· |
| J | 4762 | hCoV-19/England/ALDP-950DA7/2020\|EPI_ISL_554435\|20 | ···················· | ···················· | ····················· | ···················· | ···················· | ···················· |
| J | 4763 | hCoV-19/England/ALDP-95324B/2020\|EPI_ISL_554437\|20 | ···················· | ···················· | ····················· | ···················· | ···················· | ···················· |
| J | 4764 | hCoV-19/England/ALDP-953FF9/2020\|EPI_ISL_554438\|20 | ···················· | ···················· | ····················· | ···················· | ···················· | ···················· |
| J | 4765 | hCoV-19/England/ALDP-9509A3/2020\|EPI_ISL_554439\|20 | ···················· | ···················· | ····················· | ···················· | ···················· | ···················· |
| J | 4766 | hCoV-19/England/ALDP-95325A/2020\|EPI_ISL_554440\|20 | ···················· | ···················· | ····················· | ···················· | ···················· | ···················· |
| J | 4767 | hCoV-19/England/ALDP-95091C/2020\|EPI_ISL_554442\|20 | ···················· | ···················· | ····················· | ···················· | ···················· | ···················· |
| J | 4768 | hCoV-19/England/ALDP-953427/2020\|EPI_ISL_554443\|20 | ···················· | ···················· | ····················· | ···················· | ···················· | ···················· |
| J | 4769 | hCoV-19/England/ALDP-9558DE/2020\|EPI_ISL_554444\|20 | ···················· | ···················· | ···········T········· | ···················· | ···················· | ···················· |
| J | 4770 | hCoV-19/England/ALDP-950B16/2020\|EPI_ISL_554445\|20 | ···················· | ···················· | ····················· | ···················· | ···················· | ···················· |
| J | 4771 | hCoV-19/England/ALDP-95AF89/2020\|EPI_ISL_555667\|20 | ···················· | ···················· | ····················· | ···················· | ···················· | ···················· |
| J | 4772 | hCoV-19/England/ALDP-94DDF8/2020\|EPI_ISL_555668\|20 | ···················· | ···················· | ····················· | ···················· | ···················· | ···················· |
| J | 4773 | hCoV-19/England/ALDP-94E270/2020\|EPI_ISL_555669\|20 | ···················· | ···················· | ····················· | ···················· | ···················· | ···················· |
| J | 4774 | hCoV-19/England/ALDP-94DC91/2020\|EPI_ISL_555670\|20 | ···················· | ···················· | ····················· | ···················· | ···················· | ···················· |
| J | 4775 | hCoV-19/England/ALDP-94E261/2020\|EPI_ISL_555671\|20 | ···················· | ···················· | ····················· | ···················· | ···················· | ···················· |
| J | 4776 | hCoV-19/England/ALDP-94D97C/2020\|EPI_ISL_555672\|20 | ···················· | ···················· | ····················· | ···················· | ···················· | ···················· |
| J | 4777 | hCoV-19/England/ALDP-95AE9B/2020\|EPI_ISL_555673\|20 | ···················· | ···················· | ····················· | ···················· | ···················· | ···················· |
| J | 4778 | hCoV-19/England/ALDP-94478A/2020\|EPI_ISL_555674\|20 | ···················· | ·A·················· | ····················· | ···················· | ···················· | ···················· |
| J | 4779 | hCoV-19/England/ALDP-66613C/2020\|EPI_ISL_555675\|20 | ···················· | ···················· | ····················· | ···················· | ···················· | ···················· |
| J | 4780 | hCoV-19/England/ALDP-666390/2020\|EPI_ISL_555676\|20 | ···················· | ···················· | ····················· | ···················· | ···················· | ···················· |
| J | 4781 | hCoV-19/Wales/ALDP-95ADF8/2020\|EPI_ISL_555677\|2020 | ···················· | ···················· | ····················· | ···················· | ···················· | ···················· |

| | | | | | | | | |
|---|---|---|---|---|---|---|---|---|
| J | 4782 | hCoV-19/England/ALDP-95ABD0/2020\|EPI_ISL_555678\|20 | ···················· | ·A·················· | ····················· | ···················· | ···················· | ···················· |
| J | 4783 | hCoV-19/England/ALDP-95B128/2020\|EPI_ISL_555679\|20 | ···················· | ···················· | ····················· | ···················· | ···················· | ···················· |
| J | 4784 | hCoV-19/England/ALDP-95AD43/2020\|EPI_ISL_555680\|20 | ···················· | ·A·················· | ····················· | ···················· | ···················· | ···················· |
| J | 4785 | hCoV-19/England/ALDP-95B0EF/2020\|EPI_ISL_555681\|20 | ···················· | ···················· | ····················· | ···················· | ···················· | ···················· |
| J | 4786 | hCoV-19/England/ALDP-94DC46/2020\|EPI_ISL_555682\|20 | ···················· | ···················· | ····················· | ···················· | ···················· | ···················· |
| J | 4787 | hCoV-19/England/ALDP-95B02B/2020\|EPI_ISL_555683\|20 | ···················· | ···················· | ····················· | ···················· | ···················· | ···················· |
| J | 4788 | hCoV-19/England/ALDP-661F46/2020\|EPI_ISL_555684\|20 | ···················· | ···················· | ····················· | ···················· | ···················· | ···················· |
| J | 4789 | hCoV-19/England/ALDP-95ADDA/2020\|EPI_ISL_555685\|20 | ···················· | ···················· | ····················· | ···················· | ···················· | ···················· |
| J | 4790 | hCoV-19/England/ALDP-6661A5/2020\|EPI_ISL_555686\|20 | ···················· | ···················· | ····················· | ···················· | ···················· | ···················· |
| J | 4791 | hCoV-19/England/ALDP-661B51/2020\|EPI_ISL_555688\|20 | ···················· | ···················· | ····················· | ···················· | ···················· | ···················· |
| J | 4792 | hCoV-19/England/ALDP-94483C/2020\|EPI_ISL_555690\|20 | ···················· | ···················· | ····················· | ···················· | ···················· | ···················· |
| J | 4793 | hCoV-19/England/ALDP-65DB76/2020\|EPI_ISL_555691\|20 | ···················· | ···················· | ····················· | ···················· | ···················· | ···················· |
| J | 4794 | hCoV-19/England/ALDP-9449B1/2020\|EPI_ISL_555693\|20 | ···················· | ···················· | ····················· | ···················· | ···················· | ···················· |
| J | 4795 | hCoV-19/England/ALDP-95AF4D/2020\|EPI_ISL_555694\|20 | ···················· | ···················· | ····················· | ···················· | ···················· | ···················· |
| J | 4796 | hCoV-19/England/ALDP-95B076/2020\|EPI_ISL_555695\|20 | ···················· | ···················· | ····················· | ···················· | ···················· | ···················· |
| J | 4797 | hCoV-19/England/ALDP-94DBEF/2020\|EPI_ISL_555696\|20 | ···················· | ···················· | ····················· | ···················· | ···················· | ···················· |
| J | 4798 | hCoV-19/England/ALDP-6661E1/2020\|EPI_ISL_555697\|20 | ···················· | ···················· | ····················· | ···················· | ···················· | ···················· |
| J | 4799 | hCoV-19/England/ALDP-95B058/2020\|EPI_ISL_555698\|20 | ···················· | ···················· | ····················· | ···················· | ···················· | ···················· |
| J | 4800 | hCoV-19/England/ALDP-94490C/2020\|EPI_ISL_555699\|20 | ···················· | ·A·················· | ····················· | ···················· | ···················· | ···················· |
| J | 4801 | hCoV-19/England/ALDP-66580B/2020\|EPI_ISL_555701\|20 | ···················· | ···················· | ····················· | ···················· | ···················· | ···················· |
| J | 4802 | hCoV-19/England/ALDP-95AC19/2020\|EPI_ISL_555702\|20 | ···················· | ···················· | ····················· | ···················· | ···················· | ···················· |
| J | 4803 | hCoV-19/England/ALDP-664BD6/2020\|EPI_ISL_555704\|20 | ···················· | ···················· | ····················· | ···················· | ···················· | ···················· |
| J | 4804 | hCoV-19/England/ALDP-94DB85/2020\|EPI_ISL_555708\|20 | ···················· | ···················· | ····················· | ···················· | ···················· | ···················· |
| J | 4805 | hCoV-19/England/ALDP-952525/2020\|EPI_ISL_555709\|20 | ···················· | ···················· | ····················· | ···················· | ···················· | ···················· |
| J | 4806 | hCoV-19/England/ALDP-944BBB/2020\|EPI_ISL_555710\|20 | ···················· | ···················· | ····················· | ···················· | ···················· | ···················· |
| J | 4807 | hCoV-19/England/ALDP-952604/2020\|EPI_ISL_555711\|20 | ···················· | ···················· | ····················· | ···················· | ···················· | ···················· |
| J | 4808 | hCoV-19/England/ALDP-94DB58/2020\|EPI_ISL_555712\|20 | ···················· | ···················· | ····················· | ···················· | ···················· | ···················· |
| J | 4809 | hCoV-19/England/ALDP-95AEF5/2020\|EPI_ISL_555713\|20 | ···················· | ···················· | ····················· | ···················· | ···················· | ···················· |
| J | 4810 | hCoV-19/England/ALDP-94DB94/2020\|EPI_ISL_555715\|20 | ···················· | ···················· | ····················· | ···················· | ···················· | ···················· |
| J | 4811 | hCoV-19/England/ALDP-9449C0/2020\|EPI_ISL_555720\|20 | ···················· | ···················· | ····················· | ···················· | ···················· | ···················· |
| J | 4812 | hCoV-19/England/ALDP-9447B7/2020\|EPI_ISL_555721\|20 | ···················· | ···················· | ····················· | ···················· | ···················· | ···················· |
| J | 4813 | hCoV-19/England/ALDP-65D9C7/2020\|EPI_ISL_555722\|20 | ···················· | ···················· | ····················· | ···················· | ···················· | ···················· |
| J | 4814 | hCoV-19/Wales/ALDP-65D736/2020\|EPI_ISL_555723\|2020 | ···················· | ···················· | ····················· | ···················· | ···················· | ···················· |
| J | 4815 | hCoV-19/England/ALDP-944975/2020\|EPI_ISL_555724\|20 | ···················· | ···················· | ····················· | ···················· | ···················· | ···················· |
| J | 4816 | hCoV-19/England/ALDP-94DCDD/2020\|EPI_ISL_555725\|20 | ···················· | ···················· | ····················· | ···················· | ···················· | ···················· |
| J | 4817 | hCoV-19/Wales/ALDP-94D912/2020\|EPI_ISL_555726\|2020 | ···················· | ···················· | ····················· | ···················· | ···················· | ···················· |
| J | 4818 | hCoV-19/England/ALDP-94DC28/2020\|EPI_ISL_555727\|20 | ···················· | ···················· | ····················· | ···················· | ···················· | ···················· |
| J | 4819 | hCoV-19/England/ALDP-95ACEC/2020\|EPI_ISL_555728\|20 | ···················· | ···················· | ····················· | ···················· | ···················· | ···················· |
| J | 4820 | hCoV-19/England/ALDP-666828/2020\|EPI_ISL_555730\|20 | ···················· | ···················· | ····················· | ···················· | ···················· | ···················· |
| J | 4821 | hCoV-19/England/ALDP-95ABA3/2020\|EPI_ISL_555731\|20 | ···················· | ···················· | ····················· | ···················· | ···················· | ···················· |
| J | 4822 | hCoV-19/England/ALDP-95AF2F/2020\|EPI_ISL_555732\|20 | ···················· | ···················· | ····················· | ···················· | ···················· | ···················· |
| J | 4823 | hCoV-19/England/ALDP-665C4B/2020\|EPI_ISL_555733\|20 | ···················· | ···················· | ····················· | ···················· | ···················· | ···················· |
| J | 4824 | hCoV-19/England/ALDP-9448F0/2020\|EPI_ISL_555734\|20 | ···················· | ···················· | ····················· | ···················· | ···················· | ···················· |
| J | 4825 | hCoV-19/England/ALDP-952428/2020\|EPI_ISL_555735\|20 | ···················· | ···················· | ····················· | ···················· | ···················· | ···················· |
| J | 4826 | hCoV-19/England/ALDP-664954/2020\|EPI_ISL_555737\|20 | ···················· | ···················· | ····················· | ···················· | ···················· | ···················· |
| J | 4827 | hCoV-19/England/ALDP-9449FD/2020\|EPI_ISL_555738\|20 | ···················· | ···················· | ····················· | ···················· | ···················· | ···················· |
| J | 4828 | hCoV-19/England/ALDP-94DBFE/2020\|EPI_ISL_555739\|20 | ···················· | ···················· | ····················· | ···················· | ···················· | ···················· |
| J | 4829 | hCoV-19/England/ALDP-944896/2020\|EPI_ISL_555740\|20 | ···················· | ···················· | ····················· | ···················· | ···················· | ···················· |
| J | 4830 | hCoV-19/England/ALDP-95AC28/2020\|EPI_ISL_555741\|20 | ···················· | ···················· | ····················· | ···················· | ···················· | ···················· |
| J | 4831 | hCoV-19/England/ALDP-66A5B4/2020\|EPI_ISL_555743\|20 | ···················· | ···················· | ····················· | ···················· | ···················· | ···················· |
| J | 4832 | hCoV-19/England/ALDP-95AD9E/2020\|EPI_ISL_555744\|20 | ···················· | ···················· | ····················· | ···················· | ···················· | ···················· |
| J | 4833 | hCoV-19/England/ALDP-66AB67/2020\|EPI_ISL_555747\|20 | ···················· | ···················· | ····················· | ···················· | ···················· | ···················· |
| J | 4834 | hCoV-19/England/ALDP-95AE5F/2020\|EPI_ISL_555748\|20 | ···················· | ···················· | ····················· | ···················· | ···················· | ···················· |
| J | 4835 | hCoV-19/England/ALDP-9449EE/2020\|EPI_ISL_555750\|20 | ···················· | ···················· | ····················· | ···················· | ···················· | ···················· |
| J | 4836 | hCoV-19/England/ALDP-666381/2020\|EPI_ISL_555751\|20 | ···················· | ···················· | ····················· | ···················· | ···················· | ···················· |
| J | 4837 | hCoV-19/Wales/ALDP-94DB1C/2020\|EPI_ISL_555752\|2020 | ···················· | ···················· | ····················· | ···················· | ···················· | ···················· |
| J | 4838 | hCoV-19/England/ALDP-94473F/2020\|EPI_ISL_555753\|20 | ···················· | ···················· | ····················· | ···················· | ···················· | ···················· |
| J | 4839 | hCoV-19/England/ALDP-95ADAD/2020\|EPI_ISL_555754\|20 | ···················· | ···················· | ····················· | ···················· | ···················· | ···················· |
| J | 4840 | hCoV-19/England/ALDP-94481E/2020\|EPI_ISL_555755\|20 | ···················· | ···················· | ····················· | ···················· | ···················· | ···················· |
| J | 4841 | hCoV-19/England/ALDP-66614B/2020\|EPI_ISL_555757\|20 | ···················· | ···················· | ····················· | ···················· | ···················· | ···················· |
| J | 4842 | hCoV-19/England/ALDP-9449A2/2020\|EPI_ISL_555758\|20 | ···················· | ·A·················· | ····················· | ···················· | ···················· | ···················· |
| J | 4843 | hCoV-19/England/ALDP-94DD8F/2020\|EPI_ISL_555759\|20 | ···················· | ···················· | ····················· | ···················· | ···················· | ···················· |
| J | 4844 | hCoV-19/England/ALDP-95AEAA/2020\|EPI_ISL_555761\|20 | ···················· | ···················· | ····················· | ···················· | ···················· | ···················· |
| J | 4845 | hCoV-19/England/ALDP-95AED7/2020\|EPI_ISL_555762\|20 | ···················· | ···················· | ····················· | ···················· | ···················· | ···················· |
| J | 4846 | hCoV-19/England/ALDP-94F7F8/2020\|EPI_ISL_555764\|20 | ···················· | ···················· | ····················· | ···················· | ···················· | ···················· |
| J | 4847 | hCoV-19/Wales/ALDP-94FEE4/2020\|EPI_ISL_555765\|2020 | ···················· | ···················· | ····················· | ···················· | ···················· | ···················· |
| J | 4848 | hCoV-19/England/ALDP-94B98D/2020\|EPI_ISL_555767\|20 | ···················· | ···················· | ····················· | ···················· | ···················· | ···················· |
| J | 4849 | hCoV-19/England/ALDP-94F8AA/2020\|EPI_ISL_555768\|20 | ···················· | ···················· | ····················· | ···················· | ···················· | ···················· |
| J | 4850 | hCoV-19/England/ALDP-94B370/2020\|EPI_ISL_555770\|20 | ···················· | ···················· | ····················· | ···················· | ···················· | ···················· |
| J | 4851 | hCoV-19/England/ALDP-94B686/2020\|EPI_ISL_555771\|20 | ···················· | ···················· | ····················· | ···················· | ···················· | ···················· |
| J | 4852 | hCoV-19/England/ALDP-94F7AD/2020\|EPI_ISL_555772\|20 | ···················· | ···················· | ····················· | ···················· | ···················· | ···················· |
| J | 4853 | hCoV-19/England/ALDP-94F78F/2020\|EPI_ISL_555773\|20 | ···················· | ···················· | ····················· | ···················· | ···················· | ···················· |
| J | 4854 | hCoV-19/England/ALDP-94BBA5/2020\|EPI_ISL_555774\|20 | ···················· | ·A·················· | ····················· | ···················· | ···················· | ···················· |
| J | 4855 | hCoV-19/England/ALDP-94B89F/2020\|EPI_ISL_555776\|20 | ···················· | ···················· | ····················· | ···················· | ···················· | ···················· |
| J | 4856 | hCoV-19/England/ALDP-8BA674/2020\|EPI_ISL_556998\|20 | ·T·················· | ···················· | ····················· | ···················· | ···················· | ···················· |
| J | 4857 | hCoV-19/Wales/PHWC-16B406/2020\|EPI_ISL_560287\|2020 | ···················· | ···················· | ····················· | ···················· | ···················· | ···················· |
| J | 4858 | hCoV-19/Wales/PHWC-16B424/2020\|EPI_ISL_560288\|2020 | ···················· | ···················· | ····················· | ···················· | ···················· | ···················· |
| J | 4859 | hCoV-19/Wales/PHWC-16B49D/2020\|EPI_ISL_560292\|2020 | ···················· | ···················· | ····················· | ···················· | ···················· | ···················· |
| J | 4861 | hCoV-19/Wales/PHWC-16B4CA/2020\|EPI_ISL_560295\|2020 | ···················· | ···················· | ····················· | ···················· | ···················· | ···················· |
| J | 4862 | hCoV-19/Wales/PHWC-16B4D9/2020\|EPI_ISL_560296\|2020 | ···················· | ···················· | ····················· | ···················· | ···················· | ···················· |

| | | | | | | | | |
|---|---|---|---|---|---|---|---|---|
| J | 4864 | hCoV-19/Wales/PHWC-16B503/2020\|EPI_ISL_560299\|2020 | ···················· | ···················· | ···················· | ···················· | ···················· | ···················· |
| J | 4865 | hCoV-19/England/QEUH-9B90C1/2020\|EPI_ISL_549700\|20 | ···················· | ···················· | ···················· | ···················· | ···················· | ···················· |
| J | 4866 | hCoV-19/England/QEUH-9B851E/2020\|EPI_ISL_549701\|20 | ···················· | ···················· | ···················· | ···················· | ···················· | ···················· |
| J | 4867 | hCoV-19/England/QEUH-9B8AB5/2020\|EPI_ISL_549702\|20 | ···················· | ···················· | ···················· | ···················· | ···················· | ···················· |
| J | 4868 | hCoV-19/England/QEUH-9B8F6B/2020\|EPI_ISL_549703\|20 | ···················· | ···················· | ···················· | ···················· | ···················· | ···················· |
| J | 4869 | hCoV-19/England/QEUH-9B9182/2020\|EPI_ISL_549704\|20 | ···················· | ···················· | ···················· | ···················· | ···················· | ···················· |
| J | 4870 | hCoV-19/England/QEUH-9B99C6/2020\|EPI_ISL_549705\|20 | ········T··········· | ···················· | ···················· | ···················· | ···················· | ···················· |
| J | 4871 | hCoV-19/England/QEUH-9B897C/2020\|EPI_ISL_549707\|20 | ···················· | ·A·················· | ···················· | ···················· | ···················· | ···A················ |
| J | 4872 | hCoV-19/England/QEUH-9B8596/2020\|EPI_ISL_549708\|20 | ···················· | ···················· | ···················· | ···················· | ····T··············· | ···················· |
| J | 4873 | hCoV-19/England/QEUH-9B9A3C/2020\|EPI_ISL_549710\|20 | ···················· | ···················· | ···················· | ···················· | ···················· | ···················· |
| J | 4874 | hCoV-19/England/QEUH-9B8F89/2020\|EPI_ISL_549711\|20 | ···················· | ···················· | ···················· | ···················· | ···················· | ···················· |
| J | 4875 | hCoV-19/England/QEUH-9B9313/2020\|EPI_ISL_549712\|20 | ···················· | ···················· | ···················· | ···················· | ···················· | ···················· |
| J | 4876 | hCoV-19/England/QEUH-9B8C37/2020\|EPI_ISL_549713\|20 | ···················· | ···················· | ···················· | ···················· | ···················· | ···················· |
| J | 4877 | hCoV-19/England/QEUH-9B889D/2020\|EPI_ISL_549714\|20 | ···················· | ·A·················· | ···················· | ···················· | ···················· | ···A················ |
| J | 4878 | hCoV-19/England/QEUH-9B8B67/2020\|EPI_ISL_549715\|20 | ···················· | ···················· | ···················· | ···················· | ···················· | ···················· |
| J | 4879 | hCoV-19/England/QEUH-9B8FE3/2020\|EPI_ISL_549717\|20 | ···················· | ···················· | ···················· | ···················· | ···················· | ···················· |
| J | 4880 | hCoV-19/England/QEUH-9B9DE8/2020\|EPI_ISL_549718\|20 | ···················· | ···················· | ···················· | ···················· | ···················· | ···················· |
| J | 4881 | hCoV-19/England/QEUH-9B850F/2020\|EPI_ISL_549720\|20 | ···················· | ·A·················· | ···················· | ···················· | ···················· | ···················· |
| J | 4882 | hCoV-19/England/QEUH-9B928F/2020\|EPI_ISL_549722\|20 | ···················· | ···················· | ···················· | ···················· | ···················· | ···················· |
| J | 4883 | hCoV-19/England/QEUH-9B8C28/2020\|EPI_ISL_549723\|20 | ···················· | ···················· | ···················· | ···················· | ···················· | ···················· |
| J | 4884 | hCoV-19/England/QEUH-9B87AF/2020\|EPI_ISL_549724\|20 | ···················· | ·A·················· | ···················· | ···················· | ···················· | ···················· |
| J | 4885 | hCoV-19/England/QEUH-9B8A5B/2020\|EPI_ISL_549725\|20 | ···················· | ···················· | ···················· | ···················· | ···················· | ···················· |
| J | 4886 | hCoV-19/England/QEUH-9B901C/2020\|EPI_ISL_549727\|20 | ···················· | ···················· | ···················· | ···················· | ···················· | ···················· |
| J | 4887 | hCoV-19/England/QEUH-9B9F6A/2020\|EPI_ISL_549728\|20 | ···················· | ···················· | ···················· | ···················· | ···················· | ···················· |
| J | 4888 | hCoV-19/England/QEUH-9B8FB6/2020\|EPI_ISL_549729\|20 | ···················· | ···················· | ···················· | ···················· | ···················· | ···················· |
| J | 4889 | hCoV-19/England/QEUH-9B8C0A/2020\|EPI_ISL_549730\|20 | ···················· | ···················· | ···················· | ···················· | ···················· | ···················· |
| J | 4890 | hCoV-19/England/QEUH-9B99A8/2020\|EPI_ISL_549731\|20 | ···················· | ···················· | ···················· | ···················· | ···················· | ···················· |
| J | 4891 | hCoV-19/England/QEUH-9B9E7C/2020\|EPI_ISL_549732\|20 | ···················· | ···················· | ···················· | ···················· | ···················· | ···················· |
| J | 4892 | hCoV-19/England/QEUH-9B854B/2020\|EPI_ISL_549733\|20 | ···················· | ···················· | ···················· | ···················· | ···················· | ···················· |
| J | 4893 | hCoV-19/England/QEUH-9B9D33/2020\|EPI_ISL_549734\|20 | ···················· | ···················· | ···················· | ···················· | ···················· | ···················· |
| J | 4894 | hCoV-19/England/QEUH-9B838D/2020\|EPI_ISL_549735\|20 | ···················· | ···················· | ···················· | ···················· | ···················· | ···················· |
| J | 4895 | hCoV-19/England/QEUH-9B9252/2020\|EPI_ISL_549736\|20 | ···················· | ···················· | ···················· | ···················· | ···················· | ···················· |
| J | 4896 | hCoV-19/England/QEUH-9B8693/2020\|EPI_ISL_549737\|20 | ···················· | ···················· | ···················· | ···················· | ····T··············· | ···················· |
| J | 4897 | hCoV-19/England/QEUH-9B85B4/2020\|EPI_ISL_549738\|20 | ···················· | ···················· | ···················· | ···················· | ····T··············· | ···················· |
| J | 4898 | hCoV-19/England/QEUH-9B8F5C/2020\|EPI_ISL_549739\|20 | ···················· | ·A·················· | ···················· | ···················· | ···················· | ···················· |
| J | 4899 | hCoV-19/England/QEUH-9B9261/2020\|EPI_ISL_549740\|20 | ···················· | ···················· | ···················· | ···················· | ···················· | ···················· |
| J | 4900 | hCoV-19/England/QEUH-9B90A3/2020\|EPI_ISL_549742\|20 | ···················· | ···················· | ···················· | ···················· | ···················· | ···················· |
| J | 4901 | hCoV-19/England/QEUH-9B9999/2020\|EPI_ISL_549745\|20 | ···················· | ···················· | ···················· | ···················· | ···················· | ···················· |
| J | 4902 | hCoV-19/England/QEUH-9B9B66/2020\|EPI_ISL_549747\|20 | ···················· | ·A·················· | ···················· | ···················· | ···················· | ···················· |
| J | 4903 | hCoV-19/England/QEUH-9B8AE2/2020\|EPI_ISL_549748\|20 | ···················· | ···················· | ···················· | ···················· | ···················· | ···················· |
| J | 4904 | hCoV-19/England/QEUH-9B88BB/2020\|EPI_ISL_549749\|20 | ···················· | ···················· | ···················· | ···················· | ···················· | ···················· |
| J | 4905 | hCoV-19/England/QEUH-9B8763/2020\|EPI_ISL_549750\|20 | ···················· | ···················· | ···················· | ···················· | ···················· | ···················· |
| J | 4906 | hCoV-19/England/QEUH-9B9340/2020\|EPI_ISL_549751\|20 | ···················· | ·A·················· | ···················· | ···················· | ···················· | ···A················ |
| J | 4907 | hCoV-19/England/QEUH-9B9D60/2020\|EPI_ISL_549752\|20 | ···················· | ···················· | ···················· | ···················· | ···················· | ···················· |
| J | 4908 | hCoV-19/England/QEUH-9B848A/2020\|EPI_ISL_549753\|20 | ···················· | ···················· | ···················· | ···················· | ···················· | ···················· |
| J | 4909 | hCoV-19/England/QEUH-9B8AD3/2020\|EPI_ISL_549754\|20 | ···················· | ···················· | ···················· | ···················· | ···················· | ···················· |
| J | 4910 | hCoV-19/England/QEUH-9B8EB9/2020\|EPI_ISL_549755\|20 | ···················· | ·A·················· | ···················· | ···················· | ···················· | ···················· |
| J | 4911 | hCoV-19/England/QEUH-9B9E8B/2020\|EPI_ISL_549756\|20 | ···················· | ···················· | ···················· | ···················· | ···················· | ···················· |
| J | 4912 | hCoV-19/England/QEUH-9B847B/2020\|EPI_ISL_549757\|20 | ···················· | ···················· | ···················· | ···················· | ···················· | ···················· |
| J | 4913 | hCoV-19/England/QEUH-9B89D6/2020\|EPI_ISL_549758\|20 | ···················· | ···················· | ···················· | ···················· | ···················· | ···················· |
| J | 4914 | hCoV-19/England/QEUH-9B8569/2020\|EPI_ISL_549759\|20 | ···················· | ···················· | ···················· | ···················· | ···················· | ···················· |
| J | 4915 | hCoV-19/England/QEUH-9B8F01/2020\|EPI_ISL_549760\|20 | ···················· | ···················· | ···················· | ···················· | ···················· | ···················· |
| J | 4916 | hCoV-19/England/QEUH-9B88CA/2020\|EPI_ISL_549761\|20 | ···················· | ···················· | ···················· | ···················· | ···················· | ···················· |
| J | 4917 | hCoV-19/England/QEUH-9B8341/2020\|EPI_ISL_549762\|20 | ···················· | ·A·················· | ···················· | ···················· | ···················· | ···················· |
| J | 4918 | hCoV-19/England/QEUH-9B9146/2020\|EPI_ISL_549763\|20 | ···················· | ···················· | ···················· | ···················· | ···················· | ···················· |
| J | 4919 | hCoV-19/England/QEUH-9B8F10/2020\|EPI_ISL_549764\|20 | ···················· | ···················· | ···················· | ···················· | ···················· | ···················· |
| J | 4920 | hCoV-19/England/QEUH-9B8815/2020\|EPI_ISL_549765\|20 | ···················· | ···················· | ···················· | ···················· | ···················· | ···················· |
| J | 4921 | hCoV-19/England/QEUH-9B9304/2020\|EPI_ISL_549766\|20 | ···················· | ···················· | ···················· | ···················· | ···················· | ···················· |
| J | 4922 | hCoV-19/England/QEUH-9B9155/2020\|EPI_ISL_549767\|20 | ···················· | ···················· | ···················· | ···················· | ······T············· | ···················· |
| J | 4923 | hCoV-19/England/QEUH-9B9D51/2020\|EPI_ISL_549768\|20 | ···················· | ···················· | ···················· | ···················· | ···················· | ···················· |
| J | 4924 | hCoV-19/England/QEUH-9B91DD/2020\|EPI_ISL_549769\|20 | ···················· | ···················· | ···················· | ···················· | ······T············· | ···················· |
| J | 4925 | hCoV-19/England/QEUH-9B8D52/2020\|EPI_ISL_549773\|20 | ···················· | ···················· | ·········T·········· | ···················· | ······T············· | ···················· |
| J | 4926 | hCoV-19/England/QEUH-9B9A2D/2020\|EPI_ISL_549774\|20 | ···················· | ···················· | ···················· | ···················· | ···················· | ···················· |
| J | 4927 | hCoV-19/England/QEUH-9B9128/2020\|EPI_ISL_549776\|20 | ···················· | ···················· | ···················· | ···················· | ···················· | ···················· |
| J | 4928 | hCoV-19/England/QEUH-9B9191/2020\|EPI_ISL_549777\|20 | ···················· | ···················· | ···················· | ···················· | ···················· | ···················· |
| J | 4929 | hCoV-19/England/QEUH-9B9DD9/2020\|EPI_ISL_549778\|20 | ···················· | ···················· | ···················· | ···················· | ···················· | ···················· |
| J | 4930 | hCoV-19/England/MILK-99E0B3/2020\|EPI_ISL_550701\|20 | ···················· | ···················· | ···················· | ···················· | ···················· | ···················· |
| J | 4931 | hCoV-19/England/MILK-99CB2D/2020\|EPI_ISL_550702\|20 | ···················· | ···················· | ···················· | ···················· | ···················· | ···················· |
| J | 4932 | hCoV-19/England/MILK-99D807/2020\|EPI_ISL_550703\|20 | ···················· | ···················· | ···················· | ···················· | ···················· | ···················· |
| J | 4933 | hCoV-19/England/MILK-99D8AD/2020\|EPI_ISL_550704\|20 | ···················· | ···················· | ···················· | ···················· | ······T············· | ···················· |
| J | 4934 | hCoV-19/England/MILK-99D8BC/2020\|EPI_ISL_550705\|20 | ···················· | ···················· | ···················· | ···················· | ···················· | ···················· |
| J | 4935 | hCoV-19/England/MILK-99E5E1/2020\|EPI_ISL_550706\|20 | ···················· | ···················· | ···················· | ···················· | ···················· | ···················· |
| J | 4936 | hCoV-19/England/MILK-99DE32/2020\|EPI_ISL_550707\|20 | ···················· | ···················· | ···················· | ···················· | ···················· | ···················· |
| J | 4937 | hCoV-19/England/MILK-99E29F/2020\|EPI_ISL_550708\|20 | ···················· | ···················· | ···················· | ···················· | ···················· | ···················· |
| J | 4938 | hCoV-19/England/MILK-99C70B/2020\|EPI_ISL_550709\|20 | ···················· | ···················· | ···················· | ···················· | ······T············· | ···················· |
| J | 4939 | hCoV-19/Wales/MILK-99C5B6/2020\|EPI_ISL_550711\|2020 | ···················· | ···················· | ···················· | ···················· | ···················· | ···················· |
| J | 4940 | hCoV-19/England/MILK-99CB3C/2020\|EPI_ISL_550712\|20 | ···················· | ···················· | ···················· | ···················· | ···················· | ···················· |
| J | 4941 | hCoV-19/England/MILK-99D1FD/2020\|EPI_ISL_550713\|20 | ···················· | ···················· | ···················· | ···················· | ···················· | ···················· |
| J | 4942 | hCoV-19/England/MILK-99E52D/2020\|EPI_ISL_550714\|20 | ···················· | ···················· | ···················· | ···················· | ···················· | ···················· |
| J | 4943 | hCoV-19/England/MILK-99E11A/2020\|EPI_ISL_550715\|20 | ···················· | ···················· | ···········T········ | ···················· | ···················· | ···················· |

```
J   4944 hCoV-19/England/MILK-99DF6C/2020|EPI_ISL_550716|20   ····················  ····················  ····················  ····················  ····················  ····················
J   4945 hCoV-19/England/MILK-99DBA4/2020|EPI_ISL_550717|20   ····················  ····················  ····················  ····················  ····················  ····················
J   4946 hCoV-19/England/MILK-99DD44/2020|EPI_ISL_550718|20   ····················  ····················  ····················  ····················  ····················  ····················
J   4947 hCoV-19/England/MILK-99D88F/2020|EPI_ISL_550719|20   ····················  ····················  ····················  ····················  ····················  ····················
J   4948 hCoV-19/England/MILK-99E38D/2020|EPI_ISL_550720|20   ····················  ····················  ····················  ····················  ····················  ····················
J   4949 hCoV-19/England/MILK-99E2AE/2020|EPI_ISL_550722|20   ····················  ·A··················  ····················  ····················  ····················  ···A················
J   4950 hCoV-19/England/MILK-99C3F8/2020|EPI_ISL_550723|20   ····················  ····················  ····················  ····················  ······T·············  ····················
J   4951 hCoV-19/England/MILK-99DD53/2020|EPI_ISL_550724|20   ····················  ····················  ····················  ····················  ····················  ····················
J   4952 hCoV-19/England/MILK-99E138/2020|EPI_ISL_550725|20   ····················  ····················  ····················  ····················  ····················  ····················
J   4953 hCoV-19/Wales/MILK-99C48C/2020|EPI_ISL_550726|2020   ····················  ····················  ····················  ····················  ····················  ····················
J   4954 hCoV-19/England/MILK-99DA3E/2020|EPI_ISL_550727|20   ····················  ····················  ····················  ····················  ····················  ····················
J   4955 hCoV-19/England/MILK-99E174/2020|EPI_ISL_550728|20   ····················  ····················  ····················  ····················  ····················  ····················
J   4956 hCoV-19/Wales/MILK-99C9BA/2020|EPI_ISL_550729|2020   ····················  ····················  ····················  ····················  ····················  ····················
J   4957 hCoV-19/England/MILK-99E37E/2020|EPI_ISL_550730|20   ········T···········  ····················  ····················  ····················  ····················  ····················
J   4958 hCoV-19/England/MILK-99D1EE/2020|EPI_ISL_550731|20   ····················  ····················  ····················  ····················  ····················  ····················
J   4959 hCoV-19/England/MILK-99E183/2020|EPI_ISL_550732|20   ····················  ····················  ····················  ····················  ····················  ····················
J   4960 hCoV-19/England/MILK-99E059/2020|EPI_ISL_550733|20   ····················  ····················  ····················  ····················  ····················  ····················
J   4961 hCoV-19/England/MILK-99E5C3/2020|EPI_ISL_550734|20   ····················  ····················  ····················  ····················  ····················  ····················
J   4962 hCoV-19/England/MILK-99C5F2/2020|EPI_ISL_550735|20   ····················  ····················  ····················  ····················  ····················  ····················
J   4963 hCoV-19/England/MILK-99D782/2020|EPI_ISL_550736|20   ····················  ····················  ····················  ····················  ····················  ····················
J   4964 hCoV-19/England/MILK-99D7DD/2020|EPI_ISL_550737|20   ····················  ····················  ····················  ····················  ····················  ····················
J   4965 hCoV-19/England/MILK-99C905/2020|EPI_ISL_550738|20   ····················  ····················  ····················  ····················  ····················  ····················
J   4966 hCoV-19/England/MILK-99DC74/2020|EPI_ISL_550740|20   ····················  ····················  ····················  ····················  ····················  ····················
J   4967 hCoV-19/England/MILK-99D9E6/2020|EPI_ISL_550741|20   ····················  ····················  ····················  ····················  ····················  ····················
J   4968 hCoV-19/England/MILK-99CAF3/2020|EPI_ISL_550742|20   ····················  ····················  ····················  ····················  ····················  ····················
J   4969 hCoV-19/England/MILK-99C783/2020|EPI_ISL_550743|20   ····················  ····················  ····················  ····················  ····················  ····················
J   4970 hCoV-19/England/MILK-99DCDE/2020|EPI_ISL_550744|20   ····················  ····················  ····················  ····················  ····················  ····················
J   4971 hCoV-19/England/MILK-99D7A0/2020|EPI_ISL_550745|20   ····················  ····················  ····················  ····················  ····················  ····················
J   4972 hCoV-19/England/MILK-99D98C/2020|EPI_ISL_550746|20   ····················  ····················  ····················  ····················  ······T·············  ····················
J   4973 hCoV-19/England/MILK-99E39C/2020|EPI_ISL_550747|20   ····················  ····················  ····················  ····················  ····················  ····················
J   4974 hCoV-19/England/MILK-99DE6F/2020|EPI_ISL_550749|20   ····················  ····················  ····················  ····················  ····················  ····················
J   4975 hCoV-19/England/MILK-99C45F/2020|EPI_ISL_550758|20   ····················  ····················  ····················  ····················  ····················  ····················
J   4976 hCoV-19/England/MILK-99E2F9/2020|EPI_ISL_550760|20   ····················  ····················  ····················  ····················  ····················  ····················
J   4977 hCoV-19/England/MILK-9AA096/2020|EPI_ISL_550882|20   ····················  ·A··················  ····················  ····················  ····················  ····················
J   4978 hCoV-19/England/MILK-9AA8DA/2020|EPI_ISL_550883|20   ····················  C···················  ····················  ····················  ····················  ····················
J   4979 hCoV-19/England/MILK-9AAB86/2020|EPI_ISL_550884|20   ····················  ····················  ····················  ····················  ····················  ····················
J   4980 hCoV-19/England/MILK-9AAC65/2020|EPI_ISL_550885|20   ····················  ····················  ····················  ····················  ····················  ····················
J   4981 hCoV-19/England/MILK-9AAC38/2020|EPI_ISL_550886|20   ····················  ····················  ····················  ····················  ····················  ····················
J   4982 hCoV-19/England/MILK-9AAC74/2020|EPI_ISL_550887|20   ····················  ·A··················  ····················  ····················  ····················  ···A················
J   4983 hCoV-19/England/MILK-9AA254/2020|EPI_ISL_550888|20   ····················  ····················  ····················  ····················  ····················  ····················
J   4984 hCoV-19/England/MILK-9AAA5C/2020|EPI_ISL_550890|20   ····················  ····················  ····················  ····················  ····················  ····················
J   4985 hCoV-19/England/MILK-9AAD80/2020|EPI_ISL_550892|20   ····················  ····················  ····················  ····················  ····················  ····················
J   4986 hCoV-19/England/MILK-9AA755/2020|EPI_ISL_550893|20   ····················  ····················  ····················  ····················  ····················  ····················
J   4987 hCoV-19/England/MILK-9AA430/2020|EPI_ISL_550894|20   ····················  ····················  ····················  ····················  ····················  ····················
J   4988 hCoV-19/England/MILK-9A9862/2020|EPI_ISL_550895|20   ····················  ····················  ····················  ····················  ····················  ····················
J   4989 hCoV-19/England/MILK-9AA843/2020|EPI_ISL_550896|20   ····················  ····················  ····················  ····················  ····················  ····················
J   4990 hCoV-19/England/MILK-9AA63A/2020|EPI_ISL_550897|20   ····················  ····················  ····················  ····················  ····················  ····················
J   4991 hCoV-19/England/MILK-9AA3CA/2020|EPI_ISL_550898|20   ····················  ····················  ····················  ····················  ····················  ····················
J   4992 hCoV-19/England/MILK-9AA7CE/2020|EPI_ISL_550899|20   ····················  ····················  ····················  ····················  ····················  ····················
J   4993 hCoV-19/England/MILK-9710C5/2020|EPI_ISL_553104|20   ····················  ·A··················  ····················  ····················  ····················  ···A················
J   4994 hCoV-19/England/MILK-956AB9/2020|EPI_ISL_554290|20   ····················  ····················  ····················  ····················  ····················  ····················
J   4995 hCoV-19/England/MILK-956D56/2020|EPI_ISL_554291|20   ····················  ····················  ····················  ····················  ····················  ····················
J   4996 hCoV-19/England/ALDP-950DF2/2020|EPI_ISL_554292|20   ····················  ····················  ····················  ····················  ····················  ····················
J   4997 hCoV-19/England/MILK-9570B6/2020|EPI_ISL_554294|20   ····················  ····················  ····················  ····················  ····················  ····················
J   4998 hCoV-19/England/ALDP-950ED1/2020|EPI_ISL_554296|20   ····················  ····················  ····················  ····················  ····················  ····················
J   4999 hCoV-19/England/MILK-95705C/2020|EPI_ISL_554297|20   ····················  ····················  ····················  ····················  ····················  ····················
J   5000 hCoV-19/England/ALDP-9510DA/2020|EPI_ISL_554298|20   ····················  ····················  ····················  ····················  ····················  ····················
J   5001 hCoV-19/England/ALDP-951025/2020|EPI_ISL_554299|20   ····················  ·A··················  ····················  ····················  ····················  ····················
J   5002 hCoV-19/England/ALDP-950A55/2020|EPI_ISL_554436|20   ····················  ····················  ····················  ····················  ····················  ····················
J   5003 hCoV-19/England/ALDP-94B3E9/2020|EPI_ISL_555766|20   ····················  ····················  ····················  ····················  ····················  ····················
J   5004 hCoV-19/England/ALDP-4FA4BE/2020|EPI_ISL_559220|20   ····················  ····················  ····················  ····················  ····················  ····················
J   5005 hCoV-19/England/MILK-4BE1DD/2020|EPI_ISL_559223|20   ····················  ····················  ····················  ····················  ····················  ····················
J   5006 hCoV-19/England/ALDP-4F43A8/2020|EPI_ISL_559225|20   ····················  ····················  ····················  ····················  ····················  ····················
J   5007 hCoV-19/England/ALDP-50316F/2020|EPI_ISL_559229|20   ····················  ····················  ····················  ····················  ····················  ····················
J   5008 hCoV-19/England/ALDP-4FF808/2020|EPI_ISL_559233|20   ····················  ····················  ···R················  ····················  ····················  ····················
J   5009 hCoV-19/England/ALDP-4FF10D/2020|EPI_ISL_559234|20   ····················  ····················  ····················  ····················  ····················  ····················
J   5010 hCoV-19/England/MILK-4C16E7/2020|EPI_ISL_559236|20   ····················  ····················  ····················  ····················  ····················  ····················
J   5011 hCoV-19/England/ALDP-4F5CF7/2020|EPI_ISL_559237|20   ····················  ····················  ····················  ····················  ····················  ····················
J   5012 hCoV-19/England/ALDP-503396/2020|EPI_ISL_559238|20   ····················  ····················  ····················  ····················  ····················  ····················
J   5013 hCoV-19/England/ALDP-4F7CAA/2020|EPI_ISL_559239|20   ····················  ····················  ····················  ····················  ····················  ····················
J   5014 hCoV-19/England/MILK-4C4DC1/2020|EPI_ISL_559241|20   ····················  ····················  ····················  ····················  ····················  ····················
J   5015 hCoV-19/England/ALDP-502EF0/2020|EPI_ISL_559242|20   ····················  ····················  ····················  ····················  ····················  ····················
J   5016 hCoV-19/England/ALDP-5048F0/2020|EPI_ISL_559245|20   ····················  ····················  ····················  ····················  ····················  ····················
J   5017 hCoV-19/England/MILK-4BF35E/2020|EPI_ISL_559247|20   ····················  ····················  ····················  ····················  ····················  ····················
J   5018 hCoV-19/England/ALDP-4F551A/2020|EPI_ISL_559248|20   ····················  ····················  ····················  ····················  ····················  ····················
J   5019 hCoV-19/England/ALDP-4F7B52/2020|EPI_ISL_559250|20   ····················  ····················  ····················  ····················  ····················  ····················
J   5020 hCoV-19/England/ALDP-505DF0/2020|EPI_ISL_559251|20   ····················  ····················  ····················  ····················  ····················  ····················
J   5021 hCoV-19/England/MILK-4B325D/2020|EPI_ISL_559253|20   ····················  ····················  ····················  ····················  ····················  ····················
J   5022 hCoV-19/England/ALDP-4F9200/2020|EPI_ISL_559255|20   ····················  ····················  ····················  ····················  ····················  ····················
J   5023 hCoV-19/England/MILK-4B43A4/2020|EPI_ISL_559256|20   ····················  ····················  ····················  ····················  ····················  ····················
```

| | | | | | | | | |
|---|---|---|---|---|---|---|---|---|
| J | 5024 | hCoV-19/England/ALDP-503D7A/2020\|EPI_ISL_559257\|20 | .................... | .................... | .................... | .................... | .................... | .................... |
| J | 5025 | hCoV-19/England/ALDP-4F50DD/2020\|EPI_ISL_559258\|20 | .................... | .................... | .................... | .................... | .................... | .................... |
| J | 5026 | hCoV-19/England/MILK-4B13C5/2020\|EPI_ISL_559259\|20 | .................... | .................... | .................... | .................... | .................... | .................... |
| J | 5027 | hCoV-19/England/ALDP-4F8C6D/2020\|EPI_ISL_559260\|20 | .................... | .................... | .................... | .................... | .................... | .................... |
| J | 5028 | hCoV-19/England/ALDP-5031D8/2020\|EPI_ISL_559262\|20 | .................... | .................... | .................... | .................... | .................... | .................... |
| J | 5029 | hCoV-19/England/MILK-4B2519/2020\|EPI_ISL_559263\|20 | .................... | .................... | .................... | .................... | .................... | .................... |
| J | 5030 | hCoV-19/England/ALDP-503BF8/2020\|EPI_ISL_559269\|20 | .................... | .................... | .................... | .................... | .................... | .................... |
| J | 5031 | hCoV-19/Wales/ALDP-4F500A/2020\|EPI_ISL_559270\|2020 | .................... | .................... | .................... | .................... | .................... | .................... |
| J | 5032 | hCoV-19/England/ALDP-4F8F55/2020\|EPI_ISL_559271\|20 | .................... | .................... | .................... | .................... | .................... | .................... |
| J | 5033 | hCoV-19/England/ALDP-50631B/2020\|EPI_ISL_559272\|20 | .................... | .................... | .................... | .................... | .................... | .................... |
| J | 5034 | hCoV-19/England/ALDP-506409/2020\|EPI_ISL_559273\|20 | .................... | .................... | .................... | .................... | .................... | .................... |
| J | 5035 | hCoV-19/England/ALDP-50040F/2020\|EPI_ISL_559274\|20 | .................... | .................... | .................... | .................... | .................... | .................... |
| J | 5036 | hCoV-19/England/ALDP-4FB4F9/2020\|EPI_ISL_559276\|20 | .................... | .................... | .................... | .................... | .................... | .................... |
| J | 5037 | hCoV-19/England/MILK-4BEEB8/2020\|EPI_ISL_559277\|20 | .................... | .................... | .................... | .................... | .................... | .................... |
| J | 5038 | hCoV-19/England/MILK-4C6767/2020\|EPI_ISL_559278\|20 | .................... | .................... | .................... | .................... | .................... | .................... |
| J | 5039 | hCoV-19/England/ALDP-4F7D89/2020\|EPI_ISL_559279\|20 | .................... | .................... | .................... | .................... | .................... | .................... |
| J | 5040 | hCoV-19/England/ALDP-4FE678/2020\|EPI_ISL_559280\|20 | .................... | .................... | .................... | .................... | .................... | .................... |
| J | 5041 | hCoV-19/England/ALDP-507435/2020\|EPI_ISL_559281\|20 | .................... | .............A...... | .................... | .................... | .................... | .................... |
| J | 5042 | hCoV-19/England/MILK-4B620F/2020\|EPI_ISL_559283\|20 | .................... | .................... | .................... | .................... | .................... | .................... |
| J | 5043 | hCoV-19/England/MILK-4B1F58/2020\|EPI_ISL_559284\|20 | .................... | .................... | .................... | .................... | .................... | .................... |
| J | 5044 | hCoV-19/England/ALDP-4F909D/2020\|EPI_ISL_559285\|20 | .................... | .................... | .................... | .................... | .................... | .................... |
| J | 5045 | hCoV-19/England/ALDP-4FC1E2/2020\|EPI_ISL_559286\|20 | .................... | .................... | .................... | .................... | .................... | .................... |
| J | 5046 | hCoV-19/England/ALDP-4F40DE/2020\|EPI_ISL_559288\|20 | .................... | .................... | .................... | .................... | .................... | .................... |
| J | 5047 | hCoV-19/England/ALDP-4F5B45/2020\|EPI_ISL_559289\|20 | .................... | .................... | .................... | .................... | .................... | .................... |
| J | 5048 | hCoV-19/England/MILK-4B30BD/2020\|EPI_ISL_559290\|20 | .................... | .................... | .................... | .................... | .................... | .................... |
| J | 5049 | hCoV-19/England/ALDP-4F7EE0/2020\|EPI_ISL_559291\|20 | .................... | .................... | .................... | .................... | .................... | .................... |
| J | 5050 | hCoV-19/England/ALDP-4FBE19/2020\|EPI_ISL_559292\|20 | .................... | .................... | .................... | .................... | .................... | .................... |
| J | 5051 | hCoV-19/England/ALDP-508E81/2020\|EPI_ISL_559293\|20 | .................... | .................... | .................... | .................... | .................... | .................... |
| J | 5052 | hCoV-19/England/ALDP-4F5A0C/2020\|EPI_ISL_559295\|20 | .................... | .................... | .................... | .................... | .................... | .................... |
| J | 5053 | hCoV-19/England/ALDP-4FFF30/2020\|EPI_ISL_559297\|20 | .................... | .................... | .................... | .................... | .................... | .................... |
| J | 5054 | hCoV-19/England/ALDP-4F7044/2020\|EPI_ISL_559298\|20 | .................... | .................... | .................... | .................... | .................... | .................... |
| J | 5055 | hCoV-19/England/ALDP-49B3C1/2020\|EPI_ISL_559299\|20 | .................... | .................... | .................... | .................... | .................... | .................... |
| J | 5056 | hCoV-19/Wales/PHWC-16ADDF/2020\|EPI_ISL_560200\|2020 | .................... | .................... | .................... | .................... | .................... | .................... |
| J | 5057 | hCoV-19/Wales/PHWC-16ADEE/2020\|EPI_ISL_560201\|2020 | .................... | .................... | .................... | .................... | .................... | .................... |
| J | 5058 | hCoV-19/Wales/PHWC-16ADFD/2020\|EPI_ISL_560202\|2020 | .................... | .................... | .................... | .................... | .................... | .................... |
| J | 5059 | hCoV-19/Wales/PHWC-16AE09/2020\|EPI_ISL_560203\|2020 | .................... | .................... | .................... | .................... | .................... | .................... |
| J | 5060 | hCoV-19/Wales/PHWC-16AE18/2020\|EPI_ISL_560204\|2020 | .................... | .................... | .................... | .................... | .................... | .................... |
| J | 5061 | hCoV-19/Wales/PHWC-16AE27/2020\|EPI_ISL_560205\|2020 | .................... | .................... | .................... | .................... | .................... | .................... |
| J | 5062 | hCoV-19/Wales/PHWC-16AE36/2020\|EPI_ISL_560206\|2020 | .................... | .................... | .................... | .................... | .................... | .................... |
| J | 5063 | hCoV-19/Wales/PHWC-16AE45/2020\|EPI_ISL_560207\|2020 | .................... | .................... | .................... | .................... | .................... | .................... |
| J | 5064 | hCoV-19/Wales/PHWC-16AE54/2020\|EPI_ISL_560208\|2020 | .................... | .................... | .................... | .................... | .................... | .................... |
| J | 5065 | hCoV-19/Wales/PHWC-16AE63/2020\|EPI_ISL_560209\|2020 | .................... | .................... | .................... | .................... | .................... | .................... |
| J | 5066 | hCoV-19/Wales/PHWC-16AE72/2020\|EPI_ISL_560210\|2020 | .................... | .................... | .................... | .................... | .................... | .................... |
| J | 5067 | hCoV-19/Wales/PHWC-16AE81/2020\|EPI_ISL_560211\|2020 | .................... | .................... | .................... | .................... | .................... | .................... |
| J | 5068 | hCoV-19/Wales/PHWC-16AEAF/2020\|EPI_ISL_560213\|2020 | .................... | .................... | .................... | .................... | .................... | .................... |
| J | 5069 | hCoV-19/Wales/PHWC-16AEBE/2020\|EPI_ISL_560214\|2020 | .................... | .................... | .................... | .................... | .................... | .................... |
| J | 5070 | hCoV-19/Wales/PHWC-16AECD/2020\|EPI_ISL_560215\|2020 | .................... | .................... | .................... | .................... | .................... | .................... |
| J | 5071 | hCoV-19/Wales/PHWC-16AEDC/2020\|EPI_ISL_560216\|2020 | .................... | .................... | .................... | .................... | .................... | .................... |
| J | 5072 | hCoV-19/Wales/PHWC-16AEEB/2020\|EPI_ISL_560217\|2020 | .................... | .................... | .................... | .................... | .................... | .................... |
| J | 5073 | hCoV-19/Wales/PHWC-16AEFA/2020\|EPI_ISL_560218\|2020 | .................... | .................... | .................... | .................... | .................... | .................... |
| J | 5074 | hCoV-19/Wales/PHWC-16AF06/2020\|EPI_ISL_560219\|2020 | .................... | .................... | .................... | .................... | .................... | .................... |
| J | 5075 | hCoV-19/Wales/PHWC-16AF15/2020\|EPI_ISL_560220\|2020 | .................... | .................... | .................... | .................... | .................... | .................... |
| J | 5076 | hCoV-19/Wales/PHWC-16AF24/2020\|EPI_ISL_560221\|2020 | .................... | .................... | .................... | .................... | .................... | .................... |
| J | 5077 | hCoV-19/Wales/PHWC-16AF33/2020\|EPI_ISL_560222\|2020 | .................... | .................... | .................... | .................... | .................... | .................... |
| J | 5078 | hCoV-19/Wales/PHWC-16AF42/2020\|EPI_ISL_560223\|2020 | .................... | .................... | .................... | .................... | .................... | .................... |
| J | 5079 | hCoV-19/Wales/PHWC-16AF51/2020\|EPI_ISL_560224\|2020 | .................... | .................... | .................... | .................... | .................... | .................... |
| J | 5080 | hCoV-19/Wales/PHWC-16AF60/2020\|EPI_ISL_560225\|2020 | .................... | .................... | .................... | .................... | .................... | .................... |
| J | 5081 | hCoV-19/Wales/PHWC-16AF7F/2020\|EPI_ISL_560226\|2020 | .................... | .................... | .................... | .................... | .................... | .................... |
| J | 5082 | hCoV-19/Wales/PHWC-16AF8E/2020\|EPI_ISL_560227\|2020 | .................... | .................... | .................... | .................... | .................... | .................... |
| J | 5083 | hCoV-19/Wales/PHWC-16AFAC/2020\|EPI_ISL_560229\|2020 | .................... | .................... | .................... | .................... | .................... | .................... |
| J | 5084 | hCoV-19/Wales/PHWC-16AFF7/2020\|EPI_ISL_560230\|2020 | .................... | .................... | .................... | .................... | .................... | .................... |
| J | 5085 | hCoV-19/Wales/PHWC-16B002/2020\|EPI_ISL_560231\|2020 | .................... | .................... | .................... | .................... | .................... | .................... |
| J | 5086 | hCoV-19/Wales/PHWC-16B011/2020\|EPI_ISL_560232\|2020 | .................... | .................... | .................... | .................... | .................... | .................... |
| J | 5087 | hCoV-19/Wales/PHWC-16B020/2020\|EPI_ISL_560233\|2020 | .................... | .................... | .................... | .................... | .................... | .................... |
| J | 5088 | hCoV-19/Wales/PHWC-16B03F/2020\|EPI_ISL_560234\|2020 | .................... | .................... | .................... | .................... | .................... | .................... |
| J | 5089 | hCoV-19/Wales/PHWC-16B04E/2020\|EPI_ISL_560235\|2020 | .................... | .................... | .................... | .................... | .................... | .................... |
| J | 5090 | hCoV-19/Wales/PHWC-16B06C/2020\|EPI_ISL_560237\|2020 | .................... | .................... | .................... | .................... | .................... | .................... |
| J | 5091 | hCoV-19/Wales/PHWC-16B07B/2020\|EPI_ISL_560238\|2020 | .................... | .................... | .................... | .................... | .................... | .................... |
| J | 5093 | hCoV-19/Wales/PHWC-16B10F/2020\|EPI_ISL_560245\|2020 | .................... | .................... | .................... | .................... | .................... | .................... |
| J | 5094 | hCoV-19/Wales/PHWC-16B11E/2020\|EPI_ISL_560246\|2020 | .................... | .................... | .................... | .................... | N................... | .................... |
| J | 5095 | hCoV-19/Wales/PHWC-16B12D/2020\|EPI_ISL_560247\|2020 | .................... | .................... | .................... | .................... | .................... | .................... |
| J | 5097 | hCoV-19/Wales/PHWC-16B14B/2020\|EPI_ISL_560249\|2020 | .................... | .................... | .................... | .................... | .................... | .................... |
| J | 5098 | hCoV-19/Wales/PHWC-16B15A/2020\|EPI_ISL_560250\|2020 | .................... | .................... | .................... | .................... | .................... | .................... |
| J | 5099 | hCoV-19/Wales/PHWC-16B178/2020\|EPI_ISL_560252\|2020 | .................... | .................... | .................... | .................... | .................... | .................... |
| J | 5101 | hCoV-19/Wales/PHWC-16B1B4/2020\|EPI_ISL_560256\|2020 | .................... | .................... | .................... | .................... | .................... | .................... |
| J | 5102 | hCoV-19/Wales/PHWC-16B1C3/2020\|EPI_ISL_560257\|2020 | .................... | .................... | .................... | .................... | .................... | .................... |
| J | 5103 | hCoV-19/Wales/PHWC-16B1D2/2020\|EPI_ISL_560258\|2020 | .................... | .................... | .................... | .................... | .................... | .................... |
| J | 5105 | hCoV-19/Wales/PHWC-16B21B/2020\|EPI_ISL_560260\|2020 | .................... | .................... | .................... | .................... | .................... | .................... |
| J | 5106 | hCoV-19/Wales/PHWC-16B22A/2020\|EPI_ISL_560261\|2020 | .................... | .................... | .................... | .................... | .................... | .................... |
| J | 5107 | hCoV-19/Wales/PHWC-16B239/2020\|EPI_ISL_560262\|2020 | .................... | .................... | .................... | .................... | .................... | .................... |

| | | | | | | | | | | |
|---|---|---|---|---|---|---|---|---|---|---|
| J | 5108 | hCoV-19/Wales/PHWC-16B257/2020\|EPI_ISL_560264\|2020 | ···················· | ···················· | ···················· | ···················· | ···················· | ···················· | | |
| J | 5109 | hCoV-19/Wales/PHWC-16B275/2020\|EPI_ISL_560266\|2020 | ···················· | ···················· | ···················· | ···················· | ···················· | ···················· | | |
| J | 5110 | hCoV-19/Wales/PHWC-16B293/2020\|EPI_ISL_560267\|2020 | ···················· | ···················· | ···················· | ···················· | ···················· | ···················· | | |
| J | 5111 | hCoV-19/Wales/PHWC-16B2A2/2020\|EPI_ISL_560268\|2020 | ···················· | ···················· | ···················· | ···················· | ···················· | ···················· | | |
| J | 5112 | hCoV-19/Wales/PHWC-16B318/2020\|EPI_ISL_560273\|2020 | ···················· | ···················· | ···················· | ···················· | ···················· | ···················· | | |
| J | 5113 | hCoV-19/Wales/PHWC-16B327/2020\|EPI_ISL_560274\|2020 | ···················· | ···················· | ···················· | ···················· | ···················· | ···················· | | |
| J | 5114 | hCoV-19/Wales/PHWC-16B336/2020\|EPI_ISL_560275\|2020 | ···················· | ···················· | ···················· | ···················· | ···················· | ···················· | | |
| J | 5115 | hCoV-19/Wales/PHWC-16B345/2020\|EPI_ISL_560276\|2020 | ···················· | ···················· | ···················· | ···················· | ···················· | ···················· | | |
| J | 5116 | hCoV-19/Wales/PHWC-16B372/2020\|EPI_ISL_560279\|2020 | ···················· | ···················· | ···················· | ···················· | ···················· | ···················· | | |
| J | 5117 | hCoV-19/Wales/PHWC-16B390/2020\|EPI_ISL_560281\|2020 | ···················· | ·A·················· | ···················· | ···················· | ···················· | ··A················· | | |
| J | 5118 | hCoV-19/Wales/PHWC-16B3AF/2020\|EPI_ISL_560282\|2020 | ···················· | ···················· | ···················· | ···················· | ···················· | ···················· | | |
| J | 5119 | hCoV-19/Wales/PHWC-16B3BE/2020\|EPI_ISL_560283\|2020 | ···················· | ···················· | ···················· | ···················· | ···················· | ···················· | | |
| J | 5120 | hCoV-19/Wales/PHWC-16B3CD/2020\|EPI_ISL_560284\|2020 | ···················· | ···················· | ···················· | ···················· | NNN················· | ···················· | | 1 |
| J | 5121 | hCoV-19/Wales/MILK-99C9E7/2020\|EPI_ISL_550750\|2020 | ···················· | ···················· | ···················· | ···················· | ···················· | ···················· | | |
| J | 5122 | hCoV-19/England/MILK-99D7FB/2020\|EPI_ISL_550751\|20 | ···················· | ···················· | ···················· | ···················· | ···················· | ···················· | | |
| J | 5123 | hCoV-19/England/MILK-99DF99/2020\|EPI_ISL_550752\|20 | ···················· | ···················· | ···················· | ···················· | ···················· | ···················· | | |
| J | 5124 | hCoV-19/England/MILK-99E587/2020\|EPI_ISL_550753\|20 | ···················· | ···················· | ···················· | ···················· | ···················· | ···················· | | |
| J | 5125 | hCoV-19/England/MILK-99C747/2020\|EPI_ISL_550754\|20 | ···················· | ·A·················· | ···················· | ···················· | ···················· | ··A················· | | |
| J | 5126 | hCoV-19/England/MILK-99C5A7/2020\|EPI_ISL_550755\|20 | ···················· | ···················· | ···················· | ···················· | ···················· | ···················· | | |
| J | 5127 | hCoV-19/England/MILK-99C4F5/2020\|EPI_ISL_550757\|20 | ···················· | ···················· | ···················· | ···················· | ···················· | ···················· | | |
| J | 5128 | hCoV-19/England/MILK-99DFA8/2020\|EPI_ISL_550759\|20 | ···················· | ···················· | ···················· | ···················· | ···················· | ···················· | | |
| J | 5129 | hCoV-19/England/MILK-956B10/2020\|EPI_ISL_554204\|20 | ···················· | ···················· | ···················· | ···················· | ···················· | ···················· | | |
| J | 5130 | hCoV-19/England/MILK-956D83/2020\|EPI_ISL_554205\|20 | ···················· | ···················· | ···················· | ···················· | ···················· | ···················· | | |
| J | 5131 | hCoV-19/England/ALDP-95119B/2020\|EPI_ISL_554207\|20 | ···················· | ···················· | ···················· | ···················· | ···················· | ···················· | | |
| J | 5132 | hCoV-19/England/MILK-9572DE/2020\|EPI_ISL_554208\|20 | ···················· | ···················· | ···················· | ···················· | ···················· | ···················· | | |
| J | 5133 | hCoV-19/England/MILK-956E71/2020\|EPI_ISL_554211\|20 | ···················· | ···················· | ···················· | ···················· | ···················· | ···················· | | |
| J | 5134 | hCoV-19/England/ALDP-9515AE/2020\|EPI_ISL_554212\|20 | ···················· | ···················A | ···················· | ···················· | ···················· | ···················· | 1 | |
| J | 5135 | hCoV-19/England/MILK-957274/2020\|EPI_ISL_554214\|20 | ···················· | ······A············· | ···················· | ···················· | ···················· | ···················· | | |
| J | 5136 | hCoV-19/England/ALDP-951395/2020\|EPI_ISL_554215\|20 | ···················· | ···················· | ···················· | ···················· | ···················· | ···················· | | |
| J | 5137 | hCoV-19/England/MILK-95720B/2020\|EPI_ISL_554217\|20 | ···················· | ···················· | ···················· | ···················· | ···················· | ···················· | | |
| J | 5138 | hCoV-19/England/MILK-95798E/2020\|EPI_ISL_554218\|20 | ···················· | ···················· | ···················· | ···················· | ···················· | ···················· | | |
| J | 5139 | hCoV-19/England/ALDP-9514A1/2020\|EPI_ISL_554219\|20 | ···················· | ···················· | ···················· | ···················· | ···················· | ···················· | | |
| J | 5140 | hCoV-19/England/MILK-9578EB/2020\|EPI_ISL_554221\|20 | ···················· | ···················· | ···················· | ···················· | ···················· | ···················· | | |
| J | 5141 | hCoV-19/England/MILK-957AC7/2020\|EPI_ISL_554223\|20 | ···················· | ···················· | ···················· | ···················· | ···················· | ···················· | | |
| J | 5142 | hCoV-19/England/MILK-9579AC/2020\|EPI_ISL_554224\|20 | ···················· | ···················· | ···················· | ···················· | ···················· | ···················· | | |
| J | 5143 | hCoV-19/England/ALDP-950DE3/2020\|EPI_ISL_554225\|20 | ···················· | ···················· | ···················· | ···················· | ···················· | ···················· | | |
| J | 5144 | hCoV-19/England/MILK-957B2E/2020\|EPI_ISL_554226\|20 | ···················· | ···················· | ···················· | ···················· | ···················· | ···················· | | |
| J | 5145 | hCoV-19/England/ALDP-951070/2020\|EPI_ISL_554227\|20 | ···················· | ···················· | ···················· | ···················· | ···················· | ···················· | | |
| J | 5146 | hCoV-19/England/MILK-957599/2020\|EPI_ISL_554228\|20 | ···················· | ···················· | ···················· | ···················· | ···················· | ···················· | | |
| J | 5147 | hCoV-19/England/ALDP-950FED/2020\|EPI_ISL_554229\|20 | ···················· | ···················· | ···················· | ···················· | ···················· | ···················· | | |
| J | 5148 | hCoV-19/England/ALDP-950F38/2020\|EPI_ISL_554230\|20 | ···················· | ···················· | ···················· | ···················· | ···················· | ···················· | | |
| J | 5149 | hCoV-19/England/MILK-957A7C/2020\|EPI_ISL_554231\|20 | ···················· | ···················· | ···················· | ···················· | ···················· | ···················· | | |
| J | 5150 | hCoV-19/England/ALDP-9510BC/2020\|EPI_ISL_554233\|20 | ···················· | ···················· | ···················· | ···················· | ···················· | ···················· | | |
| J | 5151 | hCoV-19/England/ALDP-951104/2020\|EPI_ISL_554235\|20 | ···················· | ···················· | ···················· | ···················· | ···················· | ···················· | | |
| J | 5152 | hCoV-19/England/MILK-956B7A/2020\|EPI_ISL_554237\|20 | ···················· | ···················· | ···················· | ···················· | ···················· | ···················· | | |
| J | 5153 | hCoV-19/England/ALDP-9512C5/2020\|EPI_ISL_554238\|20 | ···················· | ···················· | ···················· | ···················· | ···················· | ···················· | | |
| J | 5154 | hCoV-19/England/MILK-95698F/2020\|EPI_ISL_554239\|20 | ···················· | ···················· | ···················· | ···················· | ···················· | ···················· | | |
| J | 5155 | hCoV-19/England/MILK-95713B/2020\|EPI_ISL_554240\|20 | ···················· | ···················· | ···················· | ···················· | ···················· | ···················· | | |
| J | 5156 | hCoV-19/England/MILK-958DCD/2020\|EPI_ISL_554242\|20 | ···················· | ···················· | ···················· | ···················· | ···················· | ···················· | | |
| J | 5157 | hCoV-19/England/MILK-9570E3/2020\|EPI_ISL_554243\|20 | ···················· | ···················· | ···················· | ···················· | ···················· | ···················· | | |
| J | 5158 | hCoV-19/England/MILK-957168/2020\|EPI_ISL_554244\|20 | ···················· | ···················· | ···················· | ···················· | ···················· | ···················· | | |
| J | 5159 | hCoV-19/England/MILK-956FF6/2020\|EPI_ISL_554245\|20 | ···················· | ···················· | ···················· | ···················· | ···················· | ···················· | | |
| J | 5160 | hCoV-19/England/MILK-956ECC/2020\|EPI_ISL_554246\|20 | ···················· | ···················· | ···················· | ···················· | ···················· | ···················· | | |
| J | 5161 | hCoV-19/England/MILK-956CC2/2020\|EPI_ISL_554247\|20 | ···················· | ···················· | ···················· | ···················· | ···················· | ···················· | | |
| J | 5162 | hCoV-19/England/MILK-956907/2020\|EPI_ISL_554249\|20 | ···················· | ···················· | ···················· | ···················· | ···················· | ···················· | | |
| J | 5163 | hCoV-19/England/ALDP-951474/2020\|EPI_ISL_554252\|20 | ···················· | ···················· | ···················· | ···················· | ···················· | ···················· | | |
| J | 5164 | hCoV-19/England/MILK-956A40/2020\|EPI_ISL_554253\|20 | ···················· | ···················· | ···················· | ···················· | ···················· | ···················· | | |
| J | 5165 | hCoV-19/England/MILK-956A9B/2020\|EPI_ISL_554254\|20 | ···················· | ···················· | ···················· | ···················· | ···················· | ···················· | | |
| J | 5166 | hCoV-19/England/MILK-957AB8/2020\|EPI_ISL_554255\|20 | ···················· | ···················· | ···················· | ···················· | ···················· | ···················· | | |
| J | 5167 | hCoV-19/England/MILK-95707A/2020\|EPI_ISL_554256\|20 | ···················· | ···················· | ···················· | ···················· | ···················· | ···················· | | |
| J | 5168 | hCoV-19/England/ALDP-9512D4/2020\|EPI_ISL_554257\|20 | ···················· | ·A·················· | ···················· | ···················· | ···················· | ···················· | | |
| J | 5169 | hCoV-19/England/MILK-9570D4/2020\|EPI_ISL_554258\|20 | ···················· | ···················· | ···················· | ···················· | ···················· | ···················· | | |
| J | 5170 | hCoV-19/England/ALDP-951131/2020\|EPI_ISL_554260\|20 | ···················· | ···················· | ···················· | ···················· | ···················· | ···················· | | |
| J | 5171 | hCoV-19/England/MILK-956CD1/2020\|EPI_ISL_554261\|20 | ···················· | ···················· | ···················· | ···················· | ···················· | ···················· | | |
| J | 5172 | hCoV-19/England/MILK-956D65/2020\|EPI_ISL_554262\|20 | ···················· | ···················· | ···················· | ···················· | ···················· | ···················· | | |
| J | 5173 | hCoV-19/England/ALDP-95117D/2020\|EPI_ISL_554263\|20 | ···················· | ···················· | ···················· | ···················· | ···················· | ···················· | | |
| J | 5174 | hCoV-19/England/MILK-957B97/2020\|EPI_ISL_554264\|20 | ···················· | ·A·················· | ···················· | ···················· | ···················· | ···················· | | |
| J | 5175 | hCoV-19/England/ALDP-950E77/2020\|EPI_ISL_554265\|20 | ···················· | ···················· | ···················· | ···················· | ···················· | ···················· | | |
| J | 5176 | hCoV-19/England/ALDP-9513E0/2020\|EPI_ISL_554266\|20 | ···················· | ···················· | ···················· | ···················· | ···················· | ···················· | | |
| J | 5177 | hCoV-19/England/MILK-956952/2020\|EPI_ISL_554267\|20 | ···················· | ···················· | ···················· | ···················· | ···················· | ···················· | | |
| J | 5178 | hCoV-19/England/MILK-9579F7/2020\|EPI_ISL_554269\|20 | ···················· | ···················· | ···················· | ···················· | ···················· | ···················· | | |
| J | 5179 | hCoV-19/England/ALDP-95108F/2020\|EPI_ISL_554270\|20 | ···················· | ···················· | ···················· | ···················· | ···················· | ···················· | | |
| J | 5180 | hCoV-19/England/MILK-956B98/2020\|EPI_ISL_554271\|20 | ···················· | ···················· | ···················· | ···················· | ···················· | ···················· | | |
| J | 5181 | hCoV-19/England/MILK-956925/2020\|EPI_ISL_554274\|20 | ···················· | ···················· | ···················· | ···················· | ···················· | ···················· | | |
| J | 5182 | hCoV-19/England/ALDP-951492/2020\|EPI_ISL_554275\|20 | ···················· | ···················· | ···················· | ···················· | ···················· | ···················· | | |
| J | 5183 | hCoV-19/England/MILK-9570F2/2020\|EPI_ISL_554276\|20 | ···················· | ···················· | ···················· | ···················· | ···················· | ···················· | | |
| J | 5184 | hCoV-19/England/ALDP-950FB0/2020\|EPI_ISL_554278\|20 | ···················· | ···················· | ···················· | ···················· | ···················· | ···················· | | |
| J | 5185 | hCoV-19/England/ALDP-95116E/2020\|EPI_ISL_554279\|20 | ···················· | ···················· | ···················· | ···················· | ···················· | ···················· | | |
| J | 5186 | hCoV-19/England/ALDP-951007/2020\|EPI_ISL_554280\|20 | ···················· | ···················· | ···················· | ···················· | ···················· | ···················· | | |
| J | 5187 | hCoV-19/England/ALDP-95109E/2020\|EPI_ISL_554282\|20 | ···················· | ···················· | ···················· | ···················· | ···················· | ···················· | | |

| | | | | | | | |
|---|---|---|---|---|---|---|---|
| J | 5188 hCoV-19/England/ALDP-9515CC/2020\|EPI_ISL_554283\|20 | ···················· | ···················· | ····················· | ···················· | ···················· | ···················· |
| J | 5189 hCoV-19/England/MILK-956A13/2020\|EPI_ISL_554284\|20 | ···················· | ···················· | ····················· | ···················· | ···················· | ···················· |
| J | 5190 hCoV-19/England/MILK-957195/2020\|EPI_ISL_554285\|20 | ···················· | ···················· | ····················· | ···················· | ···················· | ···················· |
| J | 5191 hCoV-19/England/MILK-9569BC/2020\|EPI_ISL_554286\|20 | ···················· | ···················· | ····················· | ···················· | ···················· | ···················· |
| J | 5192 hCoV-19/England/MILK-957B4C/2020\|EPI_ISL_554287\|20 | ···················· | ···················· | ····················· | ···················· | ···················· | ···················· |
| J | 5193 hCoV-19/England/ALDP-951140/2020\|EPI_ISL_554288\|20 | ···················· | ···················· | ····················· | ···················· | ···················· | ···················· |
| J | 5194 hCoV-19/England/MILK-956C3B/2020\|EPI_ISL_554289\|20 | ···················· | ···················· | ····················· | ···················· | ···················· | ···················· |
| J | 5195 hCoV-19/England/ALDP-94E1FB/2020\|EPI_ISL_555535\|20 | ···················· | ···················· | ····················· | ···················· | ···················· | ···················· |
| J | 5196 hCoV-19/England/ALDP-94DDDA/2020\|EPI_ISL_555537\|20 | ···················· | ···················· | ····················· | ···················· | ···················· | ···················· |
| J | 5197 hCoV-19/England/ALDP-66A8D9/2020\|EPI_ISL_555538\|20 | ···················· | ···················· | ····················· | ···················· | ···················· | ···················· |
| J | 5198 hCoV-19/England/ALDP-65EB66/2020\|EPI_ISL_555539\|20 | ···················· | ···················· | ····················· | ···················· | ···················· | ···················· |
| J | 5199 hCoV-19/England/ALDP-94DBC1/2020\|EPI_ISL_555541\|20 | ···················· | ···················· | ····················· | ···················· | ···················· | ···················· |
| J | 5200 hCoV-19/England/ALDP-94DBA3/2020\|EPI_ISL_555542\|20 | ···················· | ···················· | ····················· | ···················· | ···················· | ···················· |
| J | 5201 hCoV-19/England/ALDP-95AD70/2020\|EPI_ISL_555543\|20 | ···················· | ···················· | ····················· | ···················· | ···················· | ···················· |
| J | 5202 hCoV-19/England/ALDP-95AF01/2020\|EPI_ISL_555544\|20 | ···················· | ···················· | ····················· | ···················· | ···················· | ···················· |
| J | 5203 hCoV-19/England/ALDP-944878/2020\|EPI_ISL_555545\|20 | ···················· | ···················· | ····················· | ···················· | ···················· | ···················· |
| J | 5204 hCoV-19/England/ALDP-94DBD0/2020\|EPI_ISL_555546\|20 | ········T··········· | ···················· | ····················· | ···················· | ···················· | ···················· |
| J | 5205 hCoV-19/England/ALDP-94E1A0/2020\|EPI_ISL_555547\|20 | ···················· | ···················· | ····················· | ···················· | ···················· | ···················· |
| J | 5206 hCoV-19/England/ALDP-95ADCB/2020\|EPI_ISL_555548\|20 | ···················· | ···················· | ····················· | ···················· | ···················· | ···················· |
| J | 5207 hCoV-19/England/ALDP-95B0C1/2020\|EPI_ISL_555550\|20 | ···················· | ···················· | ····················· | ···················· | ···················· | ···················· |
| J | 5208 hCoV-19/England/ALDP-94DE40/2020\|EPI_ISL_555551\|20 | ···················· | ·A·················· | ····················· | ···················· | ···················· | ···················· |
| J | 5209 hCoV-19/England/ALDP-94D95E/2020\|EPI_ISL_555552\|20 | ···················· | ···················· | ····················· | ···················· | ···················· | ···················· |
| J | 5210 hCoV-19/England/ALDP-94DE8C/2020\|EPI_ISL_555553\|20 | ···················· | ···················· | ····················· | ···················· | ···················· | ···················· |
| J | 5211 hCoV-19/England/ALDP-664EBF/2020\|EPI_ISL_555555\|20 | ···················· | ···················· | ····················· | ···················· | ···················· | ···················· |
| J | 5212 hCoV-19/England/ALDP-944A09/2020\|EPI_ISL_555556\|20 | ···················· | ···················· | ····················· | ···················· | ···················· | ···················· |
| J | 5213 hCoV-19/England/MILK-95995F/2020\|EPI_ISL_555558\|20 | ···················· | ···················· | ····················· | ···················· | ···················· | ···················· |
| J | 5214 hCoV-19/England/ALDP-952446/2020\|EPI_ISL_555559\|20 | ···················· | ···················· | ····················· | ···················· | ···················· | ···················· |
| J | 5215 hCoV-19/Wales/ALDP-95AF10/2020\|EPI_ISL_555560\|2020 | ···················· | ···················· | ····················· | ···················· | ···················· | ···················· |
| J | 5216 hCoV-19/England/ALDP-94DD43/2020\|EPI_ISL_555561\|20 | ···················· | ···················· | ····················· | ···················· | ···················· | ···················· |
| J | 5217 hCoV-19/England/ALDP-664A06/2020\|EPI_ISL_555562\|20 | ···················· | ···················· | ····················· | ···················· | ···················· | ···················· |
| J | 5218 hCoV-19/England/ALDP-94DB0D/2020\|EPI_ISL_555563\|20 | ···················· | ···················· | ····················· | ···················· | ···················· | ···················· |
| J | 5219 hCoV-19/England/ALDP-94DD52/2020\|EPI_ISL_555564\|20 | ···················· | ···················· | ····················· | ···················· | ···················· | ···················· |
| J | 5220 hCoV-19/England/ALDP-94DDBC/2020\|EPI_ISL_555565\|20 | ···················· | ···················· | ····················· | ···················· | ···················· | ···················· |
| J | 5221 hCoV-19/England/ALDP-94DF98/2020\|EPI_ISL_555566\|20 | ···················· | ···················· | ····················· | ···················· | ···················· | ···················· |
| J | 5222 hCoV-19/England/ALDP-94E207/2020\|EPI_ISL_555567\|20 | ···················· | ···················· | ····················· | ···················· | ···················· | ···················· |
| J | 5223 hCoV-19/England/ALDP-665777/2020\|EPI_ISL_555569\|20 | ···················· | ···················· | ····················· | ···················· | ···················· | ···················· |
| J | 5224 hCoV-19/England/ALDP-95AE13/2020\|EPI_ISL_555570\|20 | ···················· | ···················· | ····················· | ···················· | ···················· | ···················· |
| J | 5225 hCoV-19/England/ALDP-95AFE3/2020\|EPI_ISL_555572\|20 | ···················· | ···················· | ····················· | ···················· | ···················· | ···················· |
| J | 5226 hCoV-19/England/ALDP-9449DF/2020\|EPI_ISL_555574\|20 | ···················· | ···················· | ····················· | ···················· | ···················· | ···················· |
| J | 5227 hCoV-19/England/ALDP-94DC55/2020\|EPI_ISL_555576\|20 | ···················· | ···················· | ····················· | ···················· | ···················· | ···················· |
| J | 5228 hCoV-19/England/ALDP-944A27/2020\|EPI_ISL_555577\|20 | ···················· | ···················· | ····················· | ···················· | ···················· | ···················· |
| J | 5229 hCoV-19/England/ALDP-94DDCB/2020\|EPI_ISL_555579\|20 | ···················· | ···················· | ····················· | ···················· | ···················· | ···················· |
| J | 5230 hCoV-19/England/ALDP-95B0FE/2020\|EPI_ISL_555580\|20 | ···················· | ···················· | ····················· | ···················· | ···················· | ···················· |
| J | 5231 hCoV-19/England/ALDP-952394/2020\|EPI_ISL_555581\|20 | ···················· | ···················· | ····················· | ···················· | ···················· | ···················· |
| J | 5232 hCoV-19/England/ALDP-95AEC8/2020\|EPI_ISL_555582\|20 | ···················· | ···················· | ····················· | ···················· | ···················· | ···················· |
| J | 5233 hCoV-19/England/ALDP-94E2CB/2020\|EPI_ISL_555583\|20 | ···················· | ···················· | ····················· | ···················· | ···················· | ···················· |
| J | 5234 hCoV-19/England/ALDP-95AFF2/2020\|EPI_ISL_555585\|20 | ···················· | ···················· | ····················· | ···················· | ···················· | ···················· |
| J | 5235 hCoV-19/England/ALDP-65CAB6/2020\|EPI_ISL_555586\|20 | ···················· | ···················· | ····················· | ···················· | ···················· | ···················· |
| J | 5236 hCoV-19/England/ALDP-95AF6B/2020\|EPI_ISL_555587\|20 | ···················· | ···················· | ····················· | ···················· | ···················· | ···················· |
| J | 5237 hCoV-19/England/ALDP-944702/2020\|EPI_ISL_555590\|20 | ···················· | ···················· | ····················· | ···················· | ···················· | ···················· |
| J | 5238 hCoV-19/England/ALDP-65E28F/2020\|EPI_ISL_555591\|20 | ···················· | ···················· | ····················· | ···················· | ···················· | ···················· |
| J | 5239 hCoV-19/England/ALDP-95AE22/2020\|EPI_ISL_555592\|20 | ···················· | ···················· | ····················· | ···················· | ···················· | ···················· |
| J | 5240 hCoV-19/England/ALDP-95B119/2020\|EPI_ISL_555593\|20 | ···················· | ···················· | ····················· | ···················· | ···················· | ···················· |
| J | 5241 hCoV-19/England/ALDP-94E1DD/2020\|EPI_ISL_555594\|20 | ···················· | ·A·················· | ····················· | ···················· | ···················· | ···················· |
| J | 5242 hCoV-19/England/ALDP-944C7C/2020\|EPI_ISL_555595\|20 | ···················· | ···················· | ····················· | ···················· | ···················· | ···················· |
| J | 5243 hCoV-19/England/ALDP-94E1CE/2020\|EPI_ISL_555596\|20 | ···················· | ···················· | ····················· | ···················· | ···················· | ···················· |
| J | 5244 hCoV-19/England/ALDP-94482D/2020\|EPI_ISL_555599\|20 | ···················· | ···················· | ····················· | ···················· | ···················· | ···················· |
| J | 5245 hCoV-19/England/ALDP-95B01C/2020\|EPI_ISL_555600\|20 | ···················· | ···················· | ····················· | ···················· | ···················· | ···················· |
| J | 5246 hCoV-19/England/ALDP-94DF7A/2020\|EPI_ISL_555601\|20 | ···················· | ···················· | ····················· | ···················· | ···················· | ···A················ |
| J | 5247 hCoV-19/England/ALDP-94E137/2020\|EPI_ISL_555602\|20 | ···················· | ···················· | ····················· | ···················· | ···················· | ···················· |
| J | 5248 hCoV-19/England/ALDP-666DDE/2020\|EPI_ISL_555603\|20 | ···················· | ···················· | ····················· | ···················· | ···················· | ···················· |
| J | 5249 hCoV-19/England/ALDP-94DD34/2020\|EPI_ISL_555604\|20 | ···················· | ···················· | ····················· | ···················· | ···················· | ···················· |
| J | 5250 hCoV-19/England/ALDP-667DCE/2020\|EPI_ISL_555605\|20 | ···················· | ···················· | ····················· | ···················· | ···················· | ···················· |
| J | 5251 hCoV-19/England/ALDP-94E234/2020\|EPI_ISL_555608\|20 | ···················· | ·A·················· | ····················· | ···················· | ···················· | ···················· |
| J | 5252 hCoV-19/England/ALDP-669D44/2020\|EPI_ISL_555610\|20 | ···················· | ···················· | ····················· | ···················· | ···················· | ···················· |
| J | 5253 hCoV-19/England/ALDP-94484B/2020\|EPI_ISL_555611\|20 | ···················· | ···················· | ····················· | ···················· | ···················· | ···················· |
| J | 5254 hCoV-19/England/ALDP-661429/2020\|EPI_ISL_555612\|20 | ···················· | ···················· | ····················· | ···················· | ···················· | ···················· |
| J | 5255 hCoV-19/England/ALDP-94DDAD/2020\|EPI_ISL_555615\|20 | ···················· | ···················· | ····················· | ···················· | ···················· | ···················· |
| J | 5256 hCoV-19/England/MILK-959931/2020\|EPI_ISL_555616\|20 | ···················· | ···················· | ····················· | ···················· | ···················· | ···················· |
| J | 5257 hCoV-19/Wales/ALDP-95AD52/2020\|EPI_ISL_555617\|2020 | ···················· | ···················· | ····················· | ···················· | ···················· | ···················· |
| J | 5258 hCoV-19/England/ALDP-9447C6/2020\|EPI_ISL_555618\|20 | ···················· | ···················· | ····················· | ···················· | ···················· | ···················· |
| J | 5259 hCoV-19/England/ALDP-94E2DA/2020\|EPI_ISL_555619\|20 | ···················· | ···················· | ····················· | ···················· | ···················· | ···················· |
| J | 5260 hCoV-19/England/ALDP-94E2BC/2020\|EPI_ISL_555620\|20 | ···················· | ···················· | ····················· | ···················· | ···················· | ···················· |
| J | 5261 hCoV-19/England/ALDP-65C9F5/2020\|EPI_ISL_555621\|20 | ···················· | ···················· | ····················· | ···················· | ···················· | ···················· |
| J | 5262 hCoV-19/England/ALDP-9524CE/2020\|EPI_ISL_555622\|20 | ···················· | ···················· | ····················· | ···················· | ···················· | ···················· |
| J | 5263 hCoV-19/England/CAMC-942036/2020\|EPI_ISL_556867\|20 | ···················· | ···················· | ····················· | ···················· | ···················· | ···················· |
| J | 5264 hCoV-19/England/CAMC-9427C8/2020\|EPI_ISL_556868\|20 | ···················· | ···················· | ····················· | ···················· | ···················· | ···················· |
| J | 5265 hCoV-19/England/CAMC-9422D6/2020\|EPI_ISL_556869\|20 | ···················· | ···················· | ····················· | ···················· | ···················· | ···················· |
| J | 5266 hCoV-19/England/CAMC-942BBD/2020\|EPI_ISL_556870\|20 | ···················· | ···················· | ····················· | ···················· | ···················· | ···················· |
| J | 5267 hCoV-19/England/CAMC-942A83/2020\|EPI_ISL_556871\|20 | ···················· | ···················· | ····················· | ···················· | ···················· | ···················· |

J 5268 hCoV-19/England/CAMC-942713/2020|EPI_ISL_556872|20 ···················· ···················· ····················· ···················· ···················· ····················
J 5269 hCoV-19/England/CAMC-9425EC/2020|EPI_ISL_556874|20 ···················· ·A·················· ····················· ···················· ···················· ····················
J 5270 hCoV-19/England/CAMC-942CAB/2020|EPI_ISL_556875|20 ···················· ···················· ····················· ···················· ···················· ····················
J 5271 hCoV-19/England/CAMC-942EB4/2020|EPI_ISL_556876|20 ···················· ···················· ····················· ···················· ···················· ····················
J 5272 hCoV-19/England/CAMC-94224F/2020|EPI_ISL_556877|20 ···················· ···················· ····················· ···················· ···················· ····················
J 5273 hCoV-19/England/CAMC-942D11/2020|EPI_ISL_556878|20 ···················· ···················· ····················· ···················· ···················· ····················
J 5274 hCoV-19/England/CAMC-94323F/2020|EPI_ISL_556879|20 ···················· ···················· ····················· ···················· ···················· ····················
J 5275 hCoV-19/England/CAMC-942E0F/2020|EPI_ISL_556880|20 ···················· ·A·················· ····················· ···················· ···················· ····················
J 5276 hCoV-19/England/CAMC-942FFD/2020|EPI_ISL_556882|20 ···················· ···················· ····················· ···················· ···················· ····················
J 5277 hCoV-19/England/CAMC-942CBA/2020|EPI_ISL_556885|20 ···················· ······A············· ····················· ···················· ···················· ····················
J 5278 hCoV-19/England/CAMC-9424EF/2020|EPI_ISL_556886|20 ···················· ···················· ····················· ···················· ···················· ····················
J 5279 hCoV-19/England/CAMC-942009/2020|EPI_ISL_556888|20 ···················· ···················· ····················· ···················· ···················· ····················
J 5280 hCoV-19/England/CAMC-942582/2020|EPI_ISL_556889|20 ···················· ···················· ····················· ···················· ···················· ····················
J 5281 hCoV-19/England/CAMC-942054/2020|EPI_ISL_556890|20 ···················· ···················· ····················· ···················· ···················· ····················
J 5282 hCoV-19/England/CAMC-941FD0/2020|EPI_ISL_556891|20 ···················· ···················· ····················· ···················· ···················· ····················
J 5283 hCoV-19/England/CAMC-942A56/2020|EPI_ISL_556892|20 ···················· ···················· ····················· ···················· ···················· ····················
J 5284 hCoV-19/England/CAMC-942CC9/2020|EPI_ISL_556893|20 ···················· ···················· ····················· ···················· ···················· ····················
J 5285 hCoV-19/England/CAMC-943017/2020|EPI_ISL_556895|20 ···················· ···················· ····················· ···················· ···················· ····················
J 5286 hCoV-19/England/CAMC-942063/2020|EPI_ISL_556896|20 ···················· ···················· ····················· ···················· ···················· ····················
J 5287 hCoV-19/England/CAMC-942A29/2020|EPI_ISL_556899|20 ···················· ···················· ····················· ···················· ···················· ····················
J 5288 hCoV-19/England/CAMC-943062/2020|EPI_ISL_556900|20 ···················· ···················· ····················· ···················· ···················· ····················
J 5289 hCoV-19/England/CAMC-942555/2020|EPI_ISL_556901|20 ···················· ···················· ····················· ···················· ···················· ····················
J 5290 hCoV-19/England/CAMC-9425CE/2020|EPI_ISL_556902|20 ···················· ···················· ····················· ···················· ···················· ····················
J 5291 hCoV-19/England/CAMC-943053/2020|EPI_ISL_556905|20 ···················· ···················· ····················· ···················· ···················· ····················
J 5292 hCoV-19/England/CAMC-943080/2020|EPI_ISL_556906|20 ···················· ···················· ····················· ···················· ···················· ····················
J 5293 hCoV-19/England/CAMC-942F2A/2020|EPI_ISL_556907|20 ···················· ···················· ····················· ···················· ···················· ····················
J 5294 hCoV-19/England/CAMC-941F3A/2020|EPI_ISL_556909|20 ···················· ···················· ····················· ···················· ···················· ····················
J 5295 hCoV-19/England/CAMC-94328A/2020|EPI_ISL_556910|20 ···················· ···················· ····················· ···················· ···················· ····················
J 5296 hCoV-19/England/CAMC-9421F7/2020|EPI_ISL_556911|20 ···················· ·A·················· ····················· ···················· ···················· ··A·················
J 5297 hCoV-19/England/CAMC-942115/2020|EPI_ISL_556912|20 ···················· ···················· ····················· ···················· ···················· ····················
J 5298 hCoV-19/England/CAMC-942AFC/2020|EPI_ISL_556913|20 ···················· ···················· ····················· ···················· ···················· ····················
J 5299 hCoV-19/England/CAMC-947F16/2020|EPI_ISL_556915|20 ···················· ···················· ····················· ···················· ···················· ····················
J 5300 hCoV-19/England/CAMC-94671F/2020|EPI_ISL_556916|20 ···················· ···················· ····················· ···················· ···················· ····················
J 5301 hCoV-19/England/CAMC-947F70/2020|EPI_ISL_556917|20 ···················· ···················· ····················· ···················· ···················· ····················
J 5302 hCoV-19/England/CAMC-9481F1/2020|EPI_ISL_556919|20 ···················· ···················· ····················· ···················· ···················· ····················
J 5303 hCoV-19/England/CAMC-94811F/2020|EPI_ISL_556920|20 ···················· ···················· ····················· ···················· ···················· ····················
J 5304 hCoV-19/England/CAMC-9452A6/2020|EPI_ISL_556921|20 ···················· ······A············· ····················· ···················· ···················· ····················
J 5305 hCoV-19/England/CAMC-9467A6/2020|EPI_ISL_556922|20 ···················· ···················· ····················· ···················· ···················· ····················
J 5306 hCoV-19/England/CAMC-947A15/2020|EPI_ISL_556923|20 ···················· ···················· ····················· ···················· ···················· ····················
J 5307 hCoV-19/England/CAMC-945965/2020|EPI_ISL_556924|20 ···················· ···················· ····················· ···················· ···················· ····················
J 5308 hCoV-19/Wales/CAMC-94771E/2020|EPI_ISL_556926|2020 ···················· ···················· ····················· ···················· ···················· ····················
J 5309 hCoV-19/England/CAMC-949925/2020|EPI_ISL_556928|20 ···················· ···················· ····················· ···················· ···················· ····················
J 5310 hCoV-19/England/CAMC-949679/2020|EPI_ISL_556930|20 ···················· ···················· ····················· ···················· ···················· ····················
J 5311 hCoV-19/England/CAMC-944EEF/2020|EPI_ISL_556931|20 ···················· ···················· ····················· ···················· ···················· ····················
J 5312 hCoV-19/England/CAMC-948917/2020|EPI_ISL_556932|20 ···················· ···················· ····················· ···················· ···················· ····················
J 5313 hCoV-19/England/CAMC-9452D3/2020|EPI_ISL_556933|20 ···················· ···················· ····················· ···················· ···················· ····················
J 5314 hCoV-19/England/CAMC-946700/2020|EPI_ISL_556934|20 ···················· ···················· ····················· ···················· ···················· ····················
J 5315 hCoV-19/England/CAMC-947E46/2020|EPI_ISL_556935|20 ···················· ···················· ····················· ···················· ···················· ····················
J 5316 hCoV-19/England/CAMC-947E55/2020|EPI_ISL_556936|20 ···················· ···················· ····················· ···················· ···················· ····················
J 5317 hCoV-19/England/CAMC-9467F1/2020|EPI_ISL_556937|20 ···················· ···················· ····················· ···················· ···················· ····················
J 5318 hCoV-19/England/CAMC-945015/2020|EPI_ISL_556938|20 ···················· ···················· ····················· ···················· ···················· ····················
J 5319 hCoV-19/England/CAMC-948276/2020|EPI_ISL_556939|20 ···················· ···················· ····················· ···················· ···················· ····················
J 5320 hCoV-19/England/CAMC-9469A0/2020|EPI_ISL_556941|20 ···················· ···················· ····················· ···················· ···················· ····················
J 5321 hCoV-19/England/CAMC-946A8F/2020|EPI_ISL_556942|20 ···················· ······A············· ····················· ···················· ···················· ····················
J 5322 hCoV-19/England/CAMC-9478B1/2020|EPI_ISL_556945|20 ···················· ···················· ····················· ···················· ···················· ····················
J 5323 hCoV-19/England/CAMC-947A24/2020|EPI_ISL_556946|20 ···················· ···················· ····················· ···················· ······T············· ····················
J 5324 hCoV-19/England/CAMC-944F28/2020|EPI_ISL_556948|20 ···················· ···················· ····················· ···················· ···················· ····················
J 5325 hCoV-19/England/CAMC-9488FC/2020|EPI_ISL_556949|20 ···················· ·A·················· ····················· ···················· ···················· ····················
J 5326 hCoV-19/England/CAMC-94572F/2020|EPI_ISL_556950|20 ···················· ···················· ····················· ···················· ···················· ····················
J 5327 hCoV-19/England/CAMC-946788/2020|EPI_ISL_556951|20 ···················· ···················· ····················· ···················· ···················· ····················
J 5328 hCoV-19/England/CAMC-9481B5/2020|EPI_ISL_556952|20 ···················· ···················· ····················· ···················· ···················· ····················
J 5329 hCoV-19/England/ALDP-50859B/2020|EPI_ISL_559180|20 ···················· ···················· ····················· ···················· ···················· ····················
J 5330 hCoV-19/England/ALDP-5089EA/2020|EPI_ISL_559182|20 ···················· ···················· ····················· ···················· ···················· ····················
J 5331 hCoV-19/England/ALDP-4F4092/2020|EPI_ISL_559183|20 ···················· ···················· ····················· ···················· ···················· ····················
J 5332 hCoV-19/England/ALDP-508935/2020|EPI_ISL_559184|20 ···················· ···················· ····················· ···················· ···················· ····················
J 5333 hCoV-19/England/ALDP-4F8D2E/2020|EPI_ISL_559185|20 ···················· ···················· ····················· ···················· ···················· ····················
J 5334 hCoV-19/England/ALDP-4F5D03/2020|EPI_ISL_559186|20 ···················· ···················· ····················· ···················· ···················· ····················
J 5335 hCoV-19/England/MILK-4BC773/2020|EPI_ISL_559187|20 ···················· ···················· ····················· ···················· ···················· ····················
J 5336 hCoV-19/England/ALDP-4FD776/2020|EPI_ISL_559188|20 ···················· ···················· ····················· ···················· ···················· ····················
J 5337 hCoV-19/England/ALDP-4FF97E/2020|EPI_ISL_559191|20 ···················· ···················· ····················· ···················· ···················· ····················
J 5338 hCoV-19/England/ALDP-4FF8BD/2020|EPI_ISL_559193|20 ···················· ···················· ····················· ···················· ···················· ····················
J 5339 hCoV-19/England/MILK-4B8470/2020|EPI_ISL_559194|20 ···················· ···················· ····················· ···················· ···················· ····················
J 5340 hCoV-19/England/MILK-4B8A23/2020|EPI_ISL_559195|20 ···················· ···················· ····················· ···················· ···················· ····················
J 5341 hCoV-19/England/ALDP-4F4F0E/2020|EPI_ISL_559197|20 ···················· ···················· ····················· ···················· ···················· ····················
J 5342 hCoV-19/England/ALDP-507DC1/2020|EPI_ISL_559198|20 ···················· ···················· ····················· ···················· ···················· ····················
J 5343 hCoV-19/England/ALDP-4FFC66/2020|EPI_ISL_559199|20 ···················· ···················· ····················· ···················· ···················· ····················
J 5344 hCoV-19/Wales/PHWC-16ABB7/2020|EPI_ISL_560166|2020 ···················· ···················· ····················· ···················· ···················· ····················
J 5345 hCoV-19/Wales/PHWC-16ABC6/2020|EPI_ISL_560167|2020 ···················· ···················· ····················· ···················· ···················· ····················
J 5346 hCoV-19/Wales/PHWC-16ABD5/2020|EPI_ISL_560168|2020 ···················· ···················· ····················· ···················· ···················· ····················
J 5347 hCoV-19/Wales/PHWC-16ABE4/2020|EPI_ISL_560169|2020 ···················· ···················· ····················· ···················· ···················· ····················

| | | | | | | | | |
|---|---|---|---|---|---|---|---|---|
| J | 5348 hCoV-19/Wales/PHWC-16ABF3/2020\|EPI_ISL_560170\|2020 | ···················· | ···················· | ····················· | ···················· | ···················· | ···················· | |
| J | 5349 hCoV-19/Wales/PHWC-16AC0F/2020\|EPI_ISL_560171\|2020 | ···················· | ···················· | ····················· | ···················· | ···················· | ···················· | |
| J | 5350 hCoV-19/Wales/PHWC-16AC1E/2020\|EPI_ISL_560172\|2020 | ···················· | ···················· | ····················· | ···················· | ···················· | ···················· | |
| J | 5351 hCoV-19/Wales/PHWC-16AC2D/2020\|EPI_ISL_560173\|2020 | ···················· | ···················· | ····················· | ···················· | ···················· | ···················· | |
| J | 5352 hCoV-19/Wales/PHWC-16AC3C/2020\|EPI_ISL_560174\|2020 | ···················· | ···················· | ····················· | ···················· | ···················· | ···················· | |
| J | 5353 hCoV-19/Wales/PHWC-16AC4B/2020\|EPI_ISL_560175\|2020 | ···················· | ···················· | ····················· | ···················· | ···················· | ···················· | |
| J | 5354 hCoV-19/Wales/PHWC-16AC5A/2020\|EPI_ISL_560176\|2020 | ···················· | ···················· | ····················· | ···················· | ···················· | ···················· | |
| J | 5355 hCoV-19/Wales/PHWC-16AC69/2020\|EPI_ISL_560177\|2020 | ···················· | ···················· | ····················· | ···················· | NNN················· | ···················· | 1 |
| J | 5356 hCoV-19/Wales/PHWC-16AC78/2020\|EPI_ISL_560178\|2020 | ···················· | ···················· | ····················· | ···················· | ···················· | ···················· | |
| J | 5357 hCoV-19/Wales/PHWC-16AC87/2020\|EPI_ISL_560179\|2020 | ···················· | ···················· | ····················· | ···················· | ···················· | ···················· | |
| J | 5358 hCoV-19/Wales/PHWC-16AC96/2020\|EPI_ISL_560180\|2020 | ···················· | ···················· | ····················· | ···················· | ···················· | ···················· | |
| J | 5359 hCoV-19/Wales/PHWC-16ACA5/2020\|EPI_ISL_560181\|2020 | ···················· | ···················· | ····················· | ···················· | ···················· | ···················· | |
| J | 5360 hCoV-19/Wales/PHWC-16ACB4/2020\|EPI_ISL_560182\|2020 | ···················· | ···················· | ····················· | ···················· | ···················· | ···················· | |
| J | 5361 hCoV-19/Wales/PHWC-16ACC3/2020\|EPI_ISL_560183\|2020 | ···················· | ···················· | ····················· | ···················· | ···················· | ···················· | |
| J | 5362 hCoV-19/Wales/PHWC-16ACE1/2020\|EPI_ISL_560185\|2020 | ···················· | ···················· | ····················· | ···················· | ···················· | ···················· | |
| J | 5363 hCoV-19/Wales/PHWC-16ACF0/2020\|EPI_ISL_560186\|2020 | ···················· | ···················· | ····················· | ···················· | ···················· | ···················· | |
| J | 5364 hCoV-19/Wales/PHWC-16AD0C/2020\|EPI_ISL_560187\|2020 | ···················· | ···················· | ····················· | ···················· | ···················· | ···················· | |
| J | 5365 hCoV-19/Wales/PHWC-16AD1B/2020\|EPI_ISL_560188\|2020 | ···················· | ···················· | ····················· | ···················· | ···················· | ···················· | |
| J | 5366 hCoV-19/Wales/PHWC-16AD2A/2020\|EPI_ISL_560189\|2020 | ···················· | ···················· | ····················· | ···················· | ···················· | ···················· | |
| J | 5367 hCoV-19/Wales/PHWC-16AD39/2020\|EPI_ISL_560190\|2020 | ···················· | ···················· | ····················· | ···················· | ···················· | ···················· | |
| J | 5368 hCoV-19/Wales/PHWC-16AD48/2020\|EPI_ISL_560191\|2020 | ···················· | ···················· | ····················· | ···················· | ···················· | ···················· | |
| J | 5369 hCoV-19/Wales/PHWC-16AD57/2020\|EPI_ISL_560192\|2020 | ···················· | ···················· | ····················· | ···················· | ···················· | ···················· | |
| J | 5370 hCoV-19/Wales/PHWC-16AD66/2020\|EPI_ISL_560193\|2020 | ···················· | ···················· | ····················· | ···················· | ···················· | ···················· | |
| J | 5371 hCoV-19/Wales/PHWC-16AD75/2020\|EPI_ISL_560194\|2020 | ···················· | ···················· | ····················· | ···················· | ···················· | ···················· | |
| J | 5372 hCoV-19/Wales/PHWC-16AD84/2020\|EPI_ISL_560195\|2020 | ···················· | ···················· | ····················· | ···················· | ···················· | ···················· | |
| J | 5373 hCoV-19/Wales/PHWC-16AD93/2020\|EPI_ISL_560196\|2020 | ···················· | ···················· | ····················· | ···················· | ···················· | ···················· | |
| J | 5374 hCoV-19/Wales/PHWC-16ADA2/2020\|EPI_ISL_560197\|2020 | ···················· | ···················· | ····················· | ···················· | ···················· | ···················· | |
| J | 5375 hCoV-19/Wales/PHWC-16ADB1/2020\|EPI_ISL_560198\|2020 | ···················· | ···················· | ····················· | ···················· | ···················· | ···················· | |
| J | 5376 hCoV-19/Wales/PHWC-16ADC0/2020\|EPI_ISL_560199\|2020 | ···················· | ···················· | ····················· | ···················· | ···················· | ···················· | |
| J | 5377 hCoV-19/England/QEUH-9B8B94/2020\|EPI_ISL_549780\|20 | ···················· | ···················· | ····················· | ···················· | ···················· | ···················· | |
| J | 5378 hCoV-19/England/QEUH-9B8E6E/2020\|EPI_ISL_549781\|20 | ···················· | ···················· | ····················· | ···················· | ···················· | ···················· | |
| J | 5379 hCoV-19/England/QEUH-9B9207/2020\|EPI_ISL_549782\|20 | ···················· | ···················· | ····················· | ···················· | ···················· | ···················· | |
| J | 5380 hCoV-19/England/QEUH-9B92AD/2020\|EPI_ISL_549783\|20 | ···················· | ···················· | ····················· | ···················· | ···················· | ···················· | |
| J | 5381 hCoV-19/England/QEUH-9B99E4/2020\|EPI_ISL_549785\|20 | ···················· | ···················· | ····················· | ···················· | ···················· | ···················· | |
| J | 5382 hCoV-19/England/QEUH-9B837E/2020\|EPI_ISL_549786\|20 | ···················· | ···················· | ····················· | ···················· | ···················· | ···················· | |
| J | 5383 hCoV-19/England/QEUH-9B8D9E/2020\|EPI_ISL_549787\|20 | ···················· | ···················· | ····················· | ···················· | ···················· | ···················· | |
| J | 5384 hCoV-19/Wales/QEUH-9B8639/2020\|EPI_ISL_549788\|2020 | ···················· | ···················· | ····················· | ···················· | ···················· | ···················· | |
| J | 5385 hCoV-19/England/QEUH-9B898B/2020\|EPI_ISL_549790\|20 | ···················· | ···················· | ····················· | ···················· | ···················· | ···················· | |
| J | 5386 hCoV-19/England/QEUH-9B85A5/2020\|EPI_ISL_549791\|20 | ···················· | ···················· | ····················· | ···················· | ···················· | ···················· | |
| J | 5387 hCoV-19/England/QEUH-9B9F00/2020\|EPI_ISL_549792\|20 | ···················· | ···················· | ····················· | ···················· | ···················· | ···················· | |
| J | 5388 hCoV-19/England/QEUH-9B8BC1/2020\|EPI_ISL_549793\|20 | ···················· | ···················· | ····················· | ···················· | ···················· | ···················· | |
| J | 5389 hCoV-19/England/QEUH-9B9A1E/2020\|EPI_ISL_549794\|20 | ···················· | ···················· | ····················· | ···················· | ···················· | ···················· | |
| J | 5390 hCoV-19/England/QEUH-9B90FE/2020\|EPI_ISL_549795\|20 | ···················· | ·A·················· | ····················· | ···················· | ···················· | ··A················· | |
| J | 5391 hCoV-19/England/QEUH-9B8CA0/2020\|EPI_ISL_549796\|20 | ···················· | ···················· | ····················· | ···················· | ···················· | ···················· | |
| J | 5392 hCoV-19/England/QEUH-9B9ED6/2020\|EPI_ISL_549797\|20 | ···················· | ···················· | ····················· | ···················· | ···················· | ···················· | |
| J | 5393 hCoV-19/England/QEUH-9B9E03/2020\|EPI_ISL_549798\|20 | ···················· | ···················· | ····················· | ···················· | ···················· | ···················· | |
| J | 5394 hCoV-19/England/QEUH-9B938C/2020\|EPI_ISL_549799\|20 | ···················· | ···················· | ····················· | ···················· | ···················· | ···················· | |
| J | 5395 hCoV-19/England/MILK-99C56B/2020\|EPI_ISL_550761\|20 | ···················· | ········A··········· | ····················· | ···················· | ······T············· | ···················· | |
| J | 5396 hCoV-19/England/MILK-99D834/2020\|EPI_ISL_550762\|20 | ···················· | ···················· | ····················· | ···················· | ···················· | ···················· | |
| J | 5397 hCoV-19/England/MILK-99C57A/2020\|EPI_ISL_550763\|20 | ···················· | ···················· | ····················· | ···················· | ···················· | ···················· | |
| J | 5398 hCoV-19/England/MILK-99C7CF/2020\|EPI_ISL_550764\|20 | ···················· | ···················· | ····················· | ···················· | ···················· | ···················· | |
| J | 5399 hCoV-19/England/MILK-99C765/2020\|EPI_ISL_550765\|20 | ···················· | ·A·················· | ····················· | ···················· | ···················· | ··A················· | |
| J | 5400 hCoV-19/England/MILK-99C5C5/2020\|EPI_ISL_550766\|20 | ···················· | ···················· | ····················· | ···················· | ···················· | ···················· | |
| J | 5401 hCoV-19/Wales/MILK-99CA7B/2020\|EPI_ISL_550767\|2020 | ···················· | ···················· | ····················· | ···················· | ···················· | ···················· | |
| J | 5402 hCoV-19/England/MILK-99DF02/2020\|EPI_ISL_550768\|20 | ···················· | ···················· | ····················· | ···················· | ···················· | ···················· | |
| J | 5403 hCoV-19/England/MILK-99DE41/2020\|EPI_ISL_550769\|20 | ···················· | ···················· | ····················· | ···················· | ···················· | ···················· | |
| J | 5404 hCoV-19/Wales/MILK-99C55C/2020\|EPI_ISL_550770\|2020 | ···················· | ···················· | ····················· | ···················· | ···················· | ···············A···· | |
| J | 5405 hCoV-19/Wales/MILK-99DAE3/2020\|EPI_ISL_550771\|2020 | ···················· | ···················· | ····················· | ···················· | ···················· | ···················· | |
| J | 5406 hCoV-19/England/MILK-99D7BF/2020\|EPI_ISL_550772\|20 | ···················· | ···················· | ····················· | ···················· | ···················· | ···················· | |
| J | 5407 hCoV-19/England/MILK-99E0E0/2020\|EPI_ISL_550773\|20 | ···················· | ···················· | ····················· | ···················· | ···················· | ···················· | |
| J | 5408 hCoV-19/England/MILK-99C64A/2020\|EPI_ISL_550774\|20 | ···················· | ···················· | ····················· | ···················· | ···················· | ···················· | |
| J | 5409 hCoV-19/England/MILK-99C61D/2020\|EPI_ISL_550775\|20 | ···················· | ···················· | ····················· | ···················· | ···················· | ···················· | |
| J | 5410 hCoV-19/England/MILK-99DD62/2020\|EPI_ISL_550776\|20 | ···················· | ···················· | ····················· | ···················· | ···················· | ···················· | |
| J | 5411 hCoV-19/England/MILK-99CAC6/2020\|EPI_ISL_550777\|20 | ···················· | ···················· | ····················· | ···················· | ···················· | ···················· | |
| J | 5412 hCoV-19/England/MILK-99E0A4/2020\|EPI_ISL_550778\|20 | ···················· | ···················· | ····················· | ···················· | ···················· | ···················· | |
| J | 5413 hCoV-19/England/MILK-99E54B/2020\|EPI_ISL_550779\|20 | ···················· | ···················· | ····················· | ···················· | ···················· | ···················· | |
| J | 5414 hCoV-19/England/MILK-99E1FC/2020\|EPI_ISL_550780\|20 | ···················· | ·A·················· | ····················· | ···················· | ···················· | ··A················· | |
| J | 5415 hCoV-19/England/MILK-99DC47/2020\|EPI_ISL_550782\|20 | ···················· | ···················· | ····················· | ···················· | ···················· | ···················· | |
| J | 5416 hCoV-19/England/MILK-99DC0B/2020\|EPI_ISL_550783\|20 | ···················· | ···················· | ····················· | ···················· | ···················· | ···················· | |
| J | 5417 hCoV-19/England/MILK-99E0FF/2020\|EPI_ISL_550784\|20 | ···················· | ···················· | ····················· | ···················· | ···················· | ···················· | |
| J | 5418 hCoV-19/England/MILK-99CB0F/2020\|EPI_ISL_550785\|20 | ···················· | ···················· | ····················· | ···················· | ···················· | ···················· | |
| J | 5419 hCoV-19/England/MILK-99E208/2020\|EPI_ISL_550786\|20 | ···················· | ···················· | ····················· | ···················· | ···················· | ···················· | |
| J | 5420 hCoV-19/England/MILK-99DC38/2020\|EPI_ISL_550787\|20 | ···················· | ···················· | ····················· | ···················· | ···················· | ···················· | |
| J | 5421 hCoV-19/England/MILK-99C6C2/2020\|EPI_ISL_550788\|20 | ···················· | ···················· | ····················· | ···················· | ···················· | ···················· | |
| J | 5422 hCoV-19/England/MILK-99C880/2020\|EPI_ISL_550789\|20 | ···················· | ···················· | ····················· | ···················· | ···················· | ···················· | |
| J | 5423 hCoV-19/England/MILK-99C817/2020\|EPI_ISL_550790\|20 | ···················· | ···················· | ····················· | ···················· | ···················· | ···················· | |
| J | 5424 hCoV-19/England/MILK-99D1DF/2020\|EPI_ISL_550791\|20 | ···················· | ···················· | ····················· | ···················· | ···················· | ···················· | |
| J | 5425 hCoV-19/England/MILK-99E3C9/2020\|EPI_ISL_550792\|20 | ···················· | ···················· | ····················· | ···················· | ···················· | ···················· | |
| J | 5426 hCoV-19/England/MILK-99C826/2020\|EPI_ISL_550793\|20 | ···················· | ···················· | ····················· | ···················· | ···················· | ···················· | |
| J | 5427 hCoV-19/England/MILK-9AA852/2020\|EPI_ISL_550794\|20 | ···················· | ···················· | ····················· | ···················· | ···················· | ···················· | |

J 5428 hCoV-19/England/MILK-9A9F30/2020|EPI_ISL_550795|20 ···················· ···················· ···················· ···················· ···················· ····················
J 5429 hCoV-19/England/MILK-9AADBD/2020|EPI_ISL_550796|20 ···················· ···················· ···················· ···················· ···················· ····················
J 5430 hCoV-19/England/MILK-9AAD44/2020|EPI_ISL_550797|20 ···················· ···················· ···················· ···················· ···················· ····················
J 5431 hCoV-19/England/MILK-9AACDE/2020|EPI_ISL_550798|20 ···················· ···················· ···················· ···················· ···················· ····················
J 5432 hCoV-19/England/MILK-9A9F7C/2020|EPI_ISL_550799|20 ···················· ···················· ···················· ···················· ···················· ····················
J 5433 hCoV-19/England/CAMC-9907F8/2020|EPI_ISL_551902|20 ···················· ·A·················· ···················· ···················· ···················· ··A·················
J 5434 hCoV-19/England/CAMC-990840/2020|EPI_ISL_551903|20 ···················· ···················· ···················· ···················· ···················· ····················
J 5435 hCoV-19/England/CAMC-990C9F/2020|EPI_ISL_551905|20 ···················· ···················· ···················· ···················· ·····T·············· ····················
J 5436 hCoV-19/England/CAMC-990AE0/2020|EPI_ISL_551906|20 ···················· ···················· ···················· ···················· ···················· ····················
J 5437 hCoV-19/England/CAMC-991162/2020|EPI_ISL_551907|20 ···················· ···················· ···················· ···················· ···················· ····················
J 5438 hCoV-19/England/CAMC-990F1E/2020|EPI_ISL_551908|20 ···················· ···················· ···················· ···················· ···················· ····················
J 5439 hCoV-19/England/CAMC-991232/2020|EPI_ISL_551909|20 ···················· ···················· ···················· ···················· ···················· ····················
J 5440 hCoV-19/England/ALDP-950E68/2020|EPI_ISL_554170|20 ···················· ···················· ···················· ···················· ···················· ····················
J 5441 hCoV-19/England/ALDP-950EFF/2020|EPI_ISL_554171|20 ···················· ···················· ···················· ···················· ···················· ····················
J 5442 hCoV-19/England/ALDP-9512B6/2020|EPI_ISL_554172|20 ···················· ···················· ···················· ···················· ···················· ····················
J 5443 hCoV-19/England/MILK-9569E9/2020|EPI_ISL_554173|20 ···················· ···················· ···················· ···················· ···················· ····················
J 5444 hCoV-19/England/ALDP-9510AD/2020|EPI_ISL_554174|20 ···················· ···················· ···················· ···················· ···················· ····················
J 5445 hCoV-19/England/ALDP-95133B/2020|EPI_ISL_554175|20 ···················· ···················· ···················· ···················· ···················· ····················
J 5446 hCoV-19/England/MILK-9572FC/2020|EPI_ISL_554178|20 ···················· ···················· ···················· ···················· ···················· ····················
J 5447 hCoV-19/England/MILK-9571C2/2020|EPI_ISL_554179|20 ···················· ···················· ···················· ···················· ···················· ····················
J 5448 hCoV-19/England/MILK-956F05/2020|EPI_ISL_554180|20 ···················· ···················· ···················· ···················· ···················· ····················
J 5449 hCoV-19/England/MILK-956EEA/2020|EPI_ISL_554181|20 ···················· ···················· ···················· ···················· ···················· ····················
J 5450 hCoV-19/England/MILK-956C68/2020|EPI_ISL_554182|20 ···················· ···················· ···················· ···················· ···················· ····················
J 5451 hCoV-19/England/MILK-956D38/2020|EPI_ISL_554184|20 ···················· ···················· ···················· ···················· ···················· ····················
J 5452 hCoV-19/England/MILK-95799D/2020|EPI_ISL_554185|20 ···················· ···················· ···················· ···················· ···················· ····················
J 5453 hCoV-19/England/ALDP-9511AA/2020|EPI_ISL_554188|20 ···················· ···················· ···················· ···················· ···················· ····················
J 5454 hCoV-19/England/ALDP-950EA4/2020|EPI_ISL_554189|20 ···················· ···················· ···················· ···················· ···················· ····················
J 5455 hCoV-19/England/MILK-956758/2020|EPI_ISL_554300|20 ···················· ·A·················· ···················· ···················· ···················· ····················
J 5456 hCoV-19/Wales/ALDP-951377/2020|EPI_ISL_554301|2020 ···················· ···················· ···················· ···················· ···················· ····················
J 5457 hCoV-19/England/MILK-957186/2020|EPI_ISL_554302|20 ···················· ···················· ···················· ···················· ···················· ····················
J 5458 hCoV-19/England/MILK-956864/2020|EPI_ISL_554304|20 ···················· ···················· ···················· ···················· ···················· ····················
J 5459 hCoV-19/England/ALDP-951113/2020|EPI_ISL_554305|20 ···················· ·A·················· ···················· ···················· ···················· ····················
J 5460 hCoV-19/England/MILK-95704D/2020|EPI_ISL_554306|20 ···················· ···················· ···················· ···················· ···················· ····················
J 5461 hCoV-19/England/MILK-9569F8/2020|EPI_ISL_554308|20 ···················· ·A·················· ···················· ···················· ···················· ····················
J 5462 hCoV-19/England/ALDP-951483/2020|EPI_ISL_554309|20 ···················· ···················· ···················· ···················· ···················· ····················
J 5463 hCoV-19/England/MILK-956EDB/2020|EPI_ISL_554310|20 ···················· ···················· ···················· ···················· ···················· ····················
J 5464 hCoV-19/England/MILK-9572B0/2020|EPI_ISL_554312|20 ···················· ···················· ···················· ···················· ···················· ····················
J 5465 hCoV-19/Wales/MILK-956855/2020|EPI_ISL_554313|2020 ···················· ···················· ···················· ···················· ···················· ····················
J 5466 hCoV-19/England/MILK-956934/2020|EPI_ISL_554314|20 ···················· ···················· ···················· ···················· ···················· ····················
J 5467 hCoV-19/England/MILK-957010/2020|EPI_ISL_554315|20 ···················· ···················· ···················· ···················· ···················· ····················
J 5468 hCoV-19/England/MILK-956DCF/2020|EPI_ISL_554317|20 ···················· ···················· ···················· ···················· ···················· ····················
J 5469 hCoV-19/England/ALDP-950F0B/2020|EPI_ISL_554318|20 ···················· ···················· ···················· ···················· ···················· ····················
J 5470 hCoV-19/England/MILK-9594E5/2020|EPI_ISL_554319|20 ···················· ···················· ···················· ···················· ···················· ····················
J 5471 hCoV-19/England/ALDP-950A64/2020|EPI_ISL_554320|20 ···················· ···················· ···················· ···················· ···················· ····················
J 5472 hCoV-19/England/ALDP-9509B2/2020|EPI_ISL_554322|20 ···················· ···················· ···················· ···················· ···················· ····················
J 5473 hCoV-19/England/ALDP-95082E/2020|EPI_ISL_554323|20 ···················· ···················· ···················· ···················· ···················· ····················
J 5474 hCoV-19/England/ALDP-953C89/2020|EPI_ISL_554324|20 ···················· ···················· ···················· ···················· ···················· ····················
J 5475 hCoV-19/England/ALDP-95AB85/2020|EPI_ISL_555623|20 ···················· ···················· ···················· ···················· ···················· ····················
J 5476 hCoV-19/England/ALDP-662DF0/2020|EPI_ISL_555624|20 ···················· ···················· ···················· ···················· ···················· ····················
J 5477 hCoV-19/England/ALDP-666266/2020|EPI_ISL_555625|20 ···················· ···················· ···················· ···················· ···················· ····················
J 5478 hCoV-19/England/ALDP-95AEE6/2020|EPI_ISL_555626|20 ···················· ···················· ···················· ···················· ···················· ····················
J 5479 hCoV-19/England/ALDP-94D921/2020|EPI_ISL_555627|20 ···················· ···················· ···················· ···················· ···················· ····················
J 5480 hCoV-19/England/ALDP-95AEB9/2020|EPI_ISL_555628|20 ···················· ···················· ···················· ···················· ···················· ····················
J 5481 hCoV-19/England/ALDP-94DB3A/2020|EPI_ISL_555629|20 ···················· ···················· ···················· ···················· ···················· ····················
J 5482 hCoV-19/England/ALDP-94492A/2020|EPI_ISL_555630|20 ···················· ···················· ···················· ···················· ···················· ····················
J 5483 hCoV-19/England/ALDP-952543/2020|EPI_ISL_555632|20 ···················· ···················· ···················· ···················· ···················· ····················
J 5484 hCoV-19/England/ALDP-94DAF1/2020|EPI_ISL_555633|20 ···················· ···················· ···················· ···················· ···················· ····················
J 5485 hCoV-19/England/ALDP-65DB2B/2020|EPI_ISL_555634|20 ···················· ···················· ···················· ···················· ···················· ····················
J 5486 hCoV-19/England/ALDP-95AF5C/2020|EPI_ISL_555635|20 ···················· ···················· ···················· ···················· ···················· ····················
J 5487 hCoV-19/England/ALDP-952613/2020|EPI_ISL_555636|20 ···················· ···················· ···················· ···················· ···················· ····················
J 5488 hCoV-19/England/ALDP-65C6FE/2020|EPI_ISL_555637|20 ···················· ···················· ···················· ···················· ···················· ····················
J 5489 hCoV-19/England/ALDP-944C8B/2020|EPI_ISL_555638|20 ···················· ···················· ···················· ···················· ···················· ····················
J 5490 hCoV-19/England/ALDP-668370/2020|EPI_ISL_555639|20 ···················· ···················· ···················· ···················· ···················· ····················
J 5491 hCoV-19/England/ALDP-95B0A3/2020|EPI_ISL_555642|20 ···················· ···················· ···················· ···················· ···················· ····················
J 5492 hCoV-19/England/ALDP-9448B4/2020|EPI_ISL_555643|20 ···················· ·A·················· ···················· ···················· ···················· ····················
J 5493 hCoV-19/England/ALDP-661CB8/2020|EPI_ISL_555644|20 ···················· ···················· ···················· ···················· ···················· ····················
J 5494 hCoV-19/England/ALDP-661DB5/2020|EPI_ISL_555645|20 ···················· ···················· ···················· ···················· ···················· ····················
J 5495 hCoV-19/England/ALDP-66217C/2020|EPI_ISL_555646|20 ···················· ···················· ···················· ···················· ···················· ····················
J 5496 hCoV-19/England/ALDP-9448A5/2020|EPI_ISL_555647|20 ···················· ···················· ···················· ···················· ···················· ····················
J 5497 hCoV-19/England/ALDP-6670B6/2020|EPI_ISL_555648|20 ···················· ···················· ···················· ···················· ···················· ····················
J 5498 hCoV-19/England/ALDP-66839E/2020|EPI_ISL_555649|20 ···················· ···················· ···················· ···················· ···················· ····················
J 5499 hCoV-19/England/ALDP-94DE7D/2020|EPI_ISL_555653|20 ···················· ·A·················· ···················· ···················· ···················· ····················
J 5500 hCoV-19/England/ALDP-6634FA/2020|EPI_ISL_555654|20 ···················· ···················· ···················· ···················· ···················· ····················
J 5501 hCoV-19/England/ALDP-9448D2/2020|EPI_ISL_555655|20 ···················· ···················· ···················· ···················· ···················· ····················
J 5502 hCoV-19/England/CAMC-944FA0/2020|EPI_ISL_556943|20 ···················· ···················· ···················· ···················· ···················· ····················
J 5503 hCoV-19/England/CAMC-9464CD/2020|EPI_ISL_556955|20 ···················· ·A·················· ···················· ···················· ···················· ····················
J 5504 hCoV-19/England/CAMC-946ABC/2020|EPI_ISL_556957|20 ···················· ······A············· ···················· ···················· ···················· ····················
J 5505 hCoV-19/England/CAMC-946A34/2020|EPI_ISL_556958|20 ···················· ···················· ···················· ···················· ···················· ····················
J 5506 hCoV-19/England/CAMC-944FBF/2020|EPI_ISL_556959|20 ···················· ···················· ···················· ···················· ···················· ····················
J 5507 hCoV-19/England/CAMC-9484AD/2020|EPI_ISL_556960|20 ···················· ···················· ···················· ···················· ···················· ····················

J 5508 hCoV-19/England/CAMC-9488ED/2020|EPI_ISL_556961|20 ···················· ···················· ···················· ···················· ···················· ····················
J 5509 hCoV-19/England/CAMC-9466B8/2020|EPI_ISL_556964|20 ···················· ···················· ···················· ···················· ···················· ····················
J 5510 hCoV-19/England/CAMC-9469FB/2020|EPI_ISL_556967|20 ···················· ···················· ···················· ···················· ···················· ····················
J 5511 hCoV-19/England/CAMC-948328/2020|EPI_ISL_556968|20 ···················· ···················· ···················· ···················· ···················· ····················
J 5512 hCoV-19/England/CAMC-946A25/2020|EPI_ISL_556969|20 ···················· ···················· ···················· ···················· ···················· ····················
J 5513 hCoV-19/England/CAMC-946B13/2020|EPI_ISL_556970|20 ···················· ···················· ···················· ···················· ···················· ····················
J 5514 hCoV-19/England/CAMC-94807C/2020|EPI_ISL_556971|20 ···················· ···················· ···················· ···················· ···················· ····················
J 5515 hCoV-19/England/CAMC-946885/2020|EPI_ISL_556973|20 ···················· ···················· ···················· ···················· ···················· ····················
J 5516 hCoV-19/England/CAMC-9464BE/2020|EPI_ISL_556974|20 ···················· ···················· ···················· ···················· ···················· ····················
J 5517 hCoV-19/England/CAMC-948258/2020|EPI_ISL_556975|20 ···················· ···················· ···················· ···················· ···················· ····················
J 5518 hCoV-19/England/CAMC-9486E3/2020|EPI_ISL_556976|20 ···················· ···················· ···················· ···················· ···················· ····················
J 5519 hCoV-19/England/CAMC-94809A/2020|EPI_ISL_556979|20 ···················· ···················· ···················· ···················· ···················· ····················
J 5520 hCoV-19/England/CAMC-948452/2020|EPI_ISL_556981|20 ···················· ·········A·········· ···················· ···················· ···················· ····················
J 5521 hCoV-19/England/CAMC-944F19/2020|EPI_ISL_556982|20 ···················· ···················· ···················· ···················· ···················· ····················
J 5522 hCoV-19/Wales/CAMC-9484E9/2020|EPI_ISL_556983|2020 ···················· ···················· ···················· ···················· ···················· ····················
J 5523 hCoV-19/England/ALDP-8B8B1D/2020|EPI_ISL_556985|20 ···················· ···················· ···················· ···················· ···················· ····················
J 5524 hCoV-19/England/ALDP-8FEFF1/2020|EPI_ISL_556986|20 ···················· ···················· ···················· ···················· ···················· ····················
J 5525 hCoV-19/England/MILK-4B2889/2020|EPI_ISL_559100|20 ···················· ···················· ···················· ···················· ···················· ····················
J 5526 hCoV-19/England/ALDP-502652/2020|EPI_ISL_559101|20 ···················· ···················· ···················· ···················· ···················· ····················
J 5527 hCoV-19/England/ALDP-50335A/2020|EPI_ISL_559102|20 ···················· ···················· ···················· ···················· ···················· ····················
J 5528 hCoV-19/England/ALDP-5087B3/2020|EPI_ISL_559105|20 ···················· ···················· ···················· ···················· ···················· ····················
J 5529 hCoV-19/England/ALDP-505482/2020|EPI_ISL_559107|20 ···················· ···················· ···················· ···················· ···················· ····················
J 5530 hCoV-19/England/ALDP-4FA490/2020|EPI_ISL_559108|20 ···················· ···················· ···················· ···················· ···················· ····················
J 5531 hCoV-19/England/ALDP-4FA4AF/2020|EPI_ISL_559109|20 ···················· ···················· ···················· ···················· ···················· ····················
J 5532 hCoV-19/England/ALDP-4F7035/2020|EPI_ISL_559110|20 ···················· ···················· ···················· ···················· ···················· ····················
J 5533 hCoV-19/England/ALDP-5000B0/2020|EPI_ISL_559111|20 ···················· ···················· ···················· ···················· ···················· ····················
J 5534 hCoV-19/England/ALDP-4FB1B6/2020|EPI_ISL_559112|20 ···················· ···················· ···················· ···················· ···················· ····················
J 5535 hCoV-19/England/ALDP-4FEB1F/2020|EPI_ISL_559113|20 ···················· ···················· ···················· ···················· ···················· ····················
J 5536 hCoV-19/England/ALDP-4F486D/2020|EPI_ISL_559114|20 ···················· ···················· ···················· ···················· ···················· ····················
J 5537 hCoV-19/England/ALDP-4F4C25/2020|EPI_ISL_559115|20 ···················· ·A·················· ···················· ···················· ···················· ····················
J 5538 hCoV-19/England/ALDP-502BDB/2020|EPI_ISL_559116|20 ···················· ···················· ···················· ···················· ···················· ····················
J 5539 hCoV-19/England/ALDP-503ED1/2020|EPI_ISL_559118|20 ···················· ···················· ···················· ···················· ···················· ····················
J 5540 hCoV-19/England/ALDP-503F0B/2020|EPI_ISL_559121|20 ···················· ···················· ···················· ···················· ···················· ····················
J 5541 hCoV-19/England/ALDP-503E2C/2020|EPI_ISL_559122|20 ···················· ···················· ···················· ···················· ···················· ····················
J 5542 hCoV-19/England/ALDP-4F7C31/2020|EPI_ISL_559123|20 ···················· ···················· ···················· ···················· ···················· ····················
J 5543 hCoV-19/England/ALDP-4F536B/2020|EPI_ISL_559127|20 ···················· ···················· ···················· ···················· ···················· ····················
J 5544 hCoV-19/England/ALDP-501538/2020|EPI_ISL_559128|20 ···················· ···················· ···················· ···················· ···················· ····················
J 5545 hCoV-19/England/ALDP-509196/2020|EPI_ISL_559130|20 ···················· ···················· ···················· ···················· ···················· ····················
J 5546 hCoV-19/England/MILK-4BBD54/2020|EPI_ISL_559132|20 ···················· ···················· ···················· ···················· ···················· ····················
J 5547 hCoV-19/England/ALDP-5074AE/2020|EPI_ISL_559134|20 ···················· ···················· ···················· ···················· ···················· ····················
J 5548 hCoV-19/England/ALDP-4F3F2D/2020|EPI_ISL_559135|20 ···················· ···················· ···················· ···················· ···················· ····················
J 5549 hCoV-19/England/MILK-4B7523/2020|EPI_ISL_559136|20 ···················· ·A·················· ···················· ···················· ···················· ····················
J 5550 hCoV-19/England/ALDP-4FC786/2020|EPI_ISL_559137|20 ···················· ···················· ···················· ···················· ···················· ····················
J 5551 hCoV-19/England/ALDP-4FF659/2020|EPI_ISL_559138|20 ···················· ···················· ···················· ···················· ···················· ····················
J 5552 hCoV-19/England/ALDP-507A60/2020|EPI_ISL_559139|20 ···················· ···················· ···················· ···················· ···················· ····················
J 5553 hCoV-19/England/ALDP-4FEEBC/2020|EPI_ISL_559140|20 ···················· ···················· ···················· ···················· ···················· ····················
J 5554 hCoV-19/England/ALDP-4FA6F4/2020|EPI_ISL_559141|20 ···················· ···················· ···················· ···················· ···················· ····················
J 5555 hCoV-19/England/ALDP-4FF8CC/2020|EPI_ISL_559144|20 ···················· ···················· ···················· ···················· ···················· ····················
J 5556 hCoV-19/England/ALDP-50287A/2020|EPI_ISL_559145|20 ···················· ···················· ···················· ···················· ···················· ····················
J 5557 hCoV-19/England/ALDP-4F9482/2020|EPI_ISL_559146|20 ···················· ···················· ···················· ···················· ···················· ····················
J 5558 hCoV-19/England/ALDP-4F8061/2020|EPI_ISL_559148|20 ···················· ···················· ···················· ···················· ···················· ····················
J 5559 hCoV-19/England/MILK-4BC4F4/2020|EPI_ISL_559149|20 ···················· ···················· ···················· ···················· ···················· ····················
J 5560 hCoV-19/England/ALDP-503B34/2020|EPI_ISL_559150|20 ···················· ···················· ···················· ···················· ···················· ····················
J 5561 hCoV-19/England/ALDP-502F48/2020|EPI_ISL_559153|20 ···················· ···················· ···················· ···················· ···················· ····················
J 5562 hCoV-19/England/MILK-4BD059/2020|EPI_ISL_559154|20 ···················· ···················· ···················· ···················· ···················· ····················
J 5563 hCoV-19/England/ALDP-508179/2020|EPI_ISL_559155|20 ···················· ···················· ···················· ···················· ···················· ····················
J 5564 hCoV-19/England/ALDP-4F7C9B/2020|EPI_ISL_559156|20 ···················· ···················· ···················· ···················· ···················· ····················
J 5565 hCoV-19/England/MILK-4C1966/2020|EPI_ISL_559157|20 ···················· ···················· ···················· ···················· ···················· ····················
J 5566 hCoV-19/England/ALDP-4FBAAC/2020|EPI_ISL_559160|20 ···················· ···················· ···················· ···················· ···················· ····················
J 5567 hCoV-19/England/ALDP-508F33/2020|EPI_ISL_559161|20 ···················· ···················· ···················· ···················· ···················· ····················
J 5568 hCoV-19/England/ALDP-4FE0B6/2020|EPI_ISL_559162|20 ···················· ···················· ···················· ···················· ···················· ····················
J 5569 hCoV-19/England/MILK-4B0F2C/2020|EPI_ISL_559163|20 ···················· ···················· ···················· ···················· ···················· ····················
J 5570 hCoV-19/England/MILK-4B9B89/2020|EPI_ISL_559164|20 ···················· ···················· ···················· ···················· ···················· ····················
J 5571 hCoV-19/England/ALDP-503CB9/2020|EPI_ISL_559165|20 ···················· ···················· ···················· ···················· ···················· ····················
J 5572 hCoV-19/England/MILK-4B13E3/2020|EPI_ISL_559167|20 ···················· ···················A ···················· ···················· ···················· ···················· 1
J 5573 hCoV-19/England/ALDP-50085E/2020|EPI_ISL_559169|20 ···················· ···················· ···················· ···················· ···················· ····················
J 5574 hCoV-19/England/ALDP-50229A/2020|EPI_ISL_559170|20 ···················· ···················· ···················· ···················· ···················· ····················
J 5575 hCoV-19/England/MILK-4C40D7/2020|EPI_ISL_559174|20 ···················· ···················· ···················· ···················· ···················· ····················
J 5576 hCoV-19/England/ALDP-502E2D/2020|EPI_ISL_559175|20 ···················· ···················· ···················· ···················· ···················· ····················
J 5577 hCoV-19/England/ALDP-50341B/2020|EPI_ISL_559177|20 ···················· ···················· ···················· ···················· ···················· ····················
J 5578 hCoV-19/England/ALDP-501ED3/2020|EPI_ISL_559178|20 ···················· ···················· ···················· ···················· ···················· ····················
J 5579 hCoV-19/England/ALDP-502F39/2020|EPI_ISL_559179|20 ···················· ···················· ···················· ···················· ···················· ····················
J 5580 hCoV-19/Wales/PHWC-16A777/2020|EPI_ISL_560100|2020 ···················· ···················· ···················· ···················· ···················· ····················
J 5581 hCoV-19/Wales/PHWC-16A786/2020|EPI_ISL_560101|2020 ···················· ···················· ···················· ···················· ···················· ····················
J 5582 hCoV-19/Wales/PHWC-16A7A4/2020|EPI_ISL_560103|2020 ···················· ···················· ···················· ···················· ···················· ····················
J 5583 hCoV-19/Wales/PHWC-16A7C2/2020|EPI_ISL_560105|2020 ···················· ···················· ···················· ···················· ···················· ····················
J 5584 hCoV-19/Wales/PHWC-16A7D1/2020|EPI_ISL_560106|2020 ···················· ···················· ···················· ···················· ···················· ····················
J 5585 hCoV-19/Wales/PHWC-16A7E0/2020|EPI_ISL_560107|2020 ···················· ···················· ···················· ···················· ···················· ····················
J 5588 hCoV-19/Wales/PHWC-16A874/2020|EPI_ISL_560115|2020 ···················· ···················· ···················· ···················· ···················· ····················
J 5589 hCoV-19/Wales/PHWC-16A883/2020|EPI_ISL_560116|2020 ···················· ···················· ···················· ···················· ···················· ····················

| | | | | | | | |
|---|---|---|---|---|---|---|---|
| J | 5591 hCoV-19/Wales/PHWC-16A8A1/2020\|EPI_ISL_560118\|2020 | ···················· | ···················· | ···················· | ···················· | ···················· | ···················· |
| J | 5592 hCoV-19/Wales/PHWC-16A8B0/2020\|EPI_ISL_560119\|2020 | ···················· | ···················· | ···················· | ···················· | ···················· | ···················· |
| J | 5593 hCoV-19/Wales/PHWC-16A8CF/2020\|EPI_ISL_560120\|2020 | ···················· | ···················· | ···················· | ···················· | ···················· | ···················· |
| J | 5594 hCoV-19/Wales/PHWC-16A8DE/2020\|EPI_ISL_560121\|2020 | ···················· | ···················· | ···················· | ···················· | ···················· | ···················· |
| J | 5595 hCoV-19/Wales/PHWC-16A8FC/2020\|EPI_ISL_560123\|2020 | ···················· | ···················· | ···················· | ···················· | ···················· | ···················· |
| J | 5597 hCoV-19/Wales/PHWC-16A935/2020\|EPI_ISL_560127\|2020 | ···················· | ···················· | ···················· | ···················· | ···················· | ···················· |
| J | 5599 hCoV-19/Wales/PHWC-16A953/2020\|EPI_ISL_560129\|2020 | ···················· | ···················· | ···················· | ···················· | ···················· | ···················· |
| J | 5600 hCoV-19/Wales/PHWC-16A962/2020\|EPI_ISL_560130\|2020 | ···················· | ···················· | ···················· | ···················· | ···················· | ···················· |
| J | 5602 hCoV-19/Wales/PHWC-16A9AE/2020\|EPI_ISL_560133\|2020 | ···················· | ···················· | ···················· | ···················· | ···················· | ···················· |
| J | 5603 hCoV-19/Wales/PHWC-16A9BD/2020\|EPI_ISL_560134\|2020 | ···················· | ···················· | ···················· | ···················· | ···················· | ···················· |
| J | 5604 hCoV-19/Wales/PHWC-16A9CC/2020\|EPI_ISL_560135\|2020 | ···················· | ···················· | ···················· | ···················· | ···················· | ···················· |
| J | 5605 hCoV-19/Wales/PHWC-16A9DB/2020\|EPI_ISL_560136\|2020 | ···················· | ···················· | ···················· | ···················· | ···················· | ···················· |
| J | 5606 hCoV-19/Wales/PHWC-16A9EA/2020\|EPI_ISL_560137\|2020 | ···················· | ···················· | ···················· | ···················· | ···················· | ···················· |
| J | 5607 hCoV-19/Wales/PHWC-16A9F9/2020\|EPI_ISL_560138\|2020 | ···················· | ···················· | ···················· | ···················· | ···················· | ···················· |
| J | 5608 hCoV-19/Wales/PHWC-16AA05/2020\|EPI_ISL_560139\|2020 | ···················· | ···················· | ···················· | ···················· | ···················· | ···················· |
| J | 5609 hCoV-19/Wales/PHWC-16AA23/2020\|EPI_ISL_560141\|2020 | ···················· | ···················· | ···················· | ···················· | ···················· | ···················· |
| J | 5610 hCoV-19/Wales/PHWC-16AA32/2020\|EPI_ISL_560142\|2020 | ···················· | ···················· | ···················· | ···················· | ···················· | ···················· |
| J | 5611 hCoV-19/Wales/PHWC-16AA41/2020\|EPI_ISL_560143\|2020 | ···················· | ···················· | ···················· | ···················· | ···················· | ···················· |
| J | 5612 hCoV-19/Wales/PHWC-16AA50/2020\|EPI_ISL_560144\|2020 | ···················· | ···················· | ···················· | ···················· | ···················· | ···················· |
| J | 5613 hCoV-19/Wales/PHWC-16AA6F/2020\|EPI_ISL_560145\|2020 | ···················· | ···················· | ···················· | ···················· | ···················· | ···················· |
| J | 5614 hCoV-19/Wales/PHWC-16AA7E/2020\|EPI_ISL_560146\|2020 | ···················· | ···················· | ···················· | ···················· | ···················· | ···················· |
| J | 5615 hCoV-19/Wales/PHWC-16AA9C/2020\|EPI_ISL_560148\|2020 | ···················· | ···················· | ···················· | ···················· | ···················· | ···················· |
| J | 5616 hCoV-19/Wales/PHWC-16AAAB/2020\|EPI_ISL_560149\|2020 | ···················· | ···················· | ···················· | ···················· | ···················· | ···················· |
| J | 5617 hCoV-19/Wales/PHWC-16AABA/2020\|EPI_ISL_560150\|2020 | ···················· | ···················· | ···················· | ···················· | ···················· | ···················· |
| J | 5618 hCoV-19/Wales/PHWC-16AAC9/2020\|EPI_ISL_560151\|2020 | ···················· | ···················· | ···················· | ···················· | ···················· | ···················· |
| J | 5619 hCoV-19/Wales/PHWC-16AAD8/2020\|EPI_ISL_560152\|2020 | ···················· | ···················· | ···················· | ···················· | ···················· | ···················· |
| J | 5620 hCoV-19/Wales/PHWC-16AAE7/2020\|EPI_ISL_560153\|2020 | ···················· | ···················· | ···················· | ···················· | ···················· | ···················· |
| J | 5621 hCoV-19/Wales/PHWC-16AAF6/2020\|EPI_ISL_560154\|2020 | ···················· | ···················· | ···················· | ···················· | ···················· | ···················· |
| J | 5622 hCoV-19/Wales/PHWC-16AB02/2020\|EPI_ISL_560155\|2020 | ···················· | ···················· | ···················· | ···················· | ···················· | ···················· |
| J | 5623 hCoV-19/Wales/PHWC-16AB11/2020\|EPI_ISL_560156\|2020 | ···················· | ···················· | ···················· | ···················· | ···················· | ···················· |
| J | 5624 hCoV-19/Wales/PHWC-16AB20/2020\|EPI_ISL_560157\|2020 | ···················· | ···················· | ···················· | ···················· | ···················· | ···················· |
| J | 5625 hCoV-19/Wales/PHWC-16AB3F/2020\|EPI_ISL_560158\|2020 | ···················· | ···················· | ···················· | ···················· | ···················· | ···················· |
| J | 5626 hCoV-19/Wales/PHWC-16AB4E/2020\|EPI_ISL_560159\|2020 | ···················· | ···················· | ···················· | ···················· | ···················· | ···················· |
| J | 5627 hCoV-19/Wales/PHWC-16AB5D/2020\|EPI_ISL_560160\|2020 | ···················· | ···················· | ···················· | ···················· | ···················· | ···················· |
| J | 5628 hCoV-19/Wales/PHWC-16AB6C/2020\|EPI_ISL_560161\|2020 | ···················· | ···················· | ···················· | ···················· | ···················· | ···················· |
| J | 5629 hCoV-19/Wales/PHWC-16AB7B/2020\|EPI_ISL_560162\|2020 | ···················· | ···················· | ···················· | ···················· | ···················· | ···················· |
| J | 5630 hCoV-19/Wales/PHWC-16AB8A/2020\|EPI_ISL_560163\|2020 | ···················· | ···················· | ···················· | ···················· | ···················· | ···················· |
| J | 5631 hCoV-19/Wales/PHWC-16AB99/2020\|EPI_ISL_560164\|2020 | ···················· | ···················· | ···················· | ···················· | ···················· | ···················· |
| J | 5632 hCoV-19/Wales/PHWC-16ABA8/2020\|EPI_ISL_560165\|2020 | ···················· | ···················· | ···················· | ···················· | ···················· | ···················· |
| J | 5633 hCoV-19/England/QEUH-9B9EE5/2020\|EPI_ISL_549601\|20 | ···················· | ···················· | ···················· | ···················· | ···················· | ···················· |
| J | 5634 hCoV-19/England/QEUH-9B862A/2020\|EPI_ISL_549602\|20 | ···················· | ···················· | ···················· | ···················· | ···················· | ···················· |
| J | 5635 hCoV-19/England/QEUH-9B839C/2020\|EPI_ISL_549603\|20 | ···················· | ···················· | ···················· | ···················· | ···················· | ···················· |
| J | 5636 hCoV-19/England/QEUH-9B8D16/2020\|EPI_ISL_549605\|20 | ···················· | ···················· | ···················· | ···················· | ···················· | ···················· |
| J | 5637 hCoV-19/England/QEUH-9B9B2A/2020\|EPI_ISL_549606\|20 | ···················· | ···················· | ···················· | ···················· | ···················· | ···················· |
| J | 5638 hCoV-19/England/QEUH-9B852D/2020\|EPI_ISL_549607\|20 | ···················· | ···················· | ···················· | ···················· | ···················· | ···················· |
| J | 5639 hCoV-19/England/QEUH-9B8DDA/2020\|EPI_ISL_549608\|20 | ···················· | ···················· | ···················· | ···················· | ···················· | ···················· |
| J | 5640 hCoV-19/England/QEUH-9B8DF8/2020\|EPI_ISL_549609\|20 | ···················· | ···················· | ···················· | ···················· | ···················· | ···················· |
| J | 5641 hCoV-19/England/QEUH-9B8727/2020\|EPI_ISL_549610\|20 | ···················· | ···················· | ···················· | ···················· | ···················· | ···················· |
| J | 5642 hCoV-19/England/QEUH-9B8FF2/2020\|EPI_ISL_549611\|20 | ···················· | ···················· | ···················· | ···················· | ···················· | ···················· |
| J | 5643 hCoV-19/England/QEUH-9B998A/2020\|EPI_ISL_549612\|20 | ···················· | ···················· | ···················· | ···················· | ···················· | ···················· |
| J | 5644 hCoV-19/England/QEUH-9B90B2/2020\|EPI_ISL_549613\|20 | ···················· | ···················· | ···················· | ···················· | ···················· | ···················· |
| J | 5645 hCoV-19/England/QEUH-9B8903/2020\|EPI_ISL_549614\|20 | ···················· | ·A·················· | ···················· | ···················· | ···················· | ···A················ |
| J | 5646 hCoV-19/England/QEUH-9B8AF1/2020\|EPI_ISL_549616\|20 | ···················· | ···················· | ···················· | ···················· | ···················· | ···················· |
| J | 5647 hCoV-19/England/QEUH-9B8587/2020\|EPI_ISL_549617\|20 | ···················· | ·A·················· | ···················· | ···················· | ···················· | ···················· |
| J | 5648 hCoV-19/England/QEUH-9B8DCB/2020\|EPI_ISL_549618\|20 | ···················· | ···················· | ···················· | ···················· | ······T············· | ···················· |
| J | 5649 hCoV-19/England/QEUH-9B8C73/2020\|EPI_ISL_549619\|20 | ···················· | ·A·······A·········· | ···················· | ···················· | ···················· | ···················· |
| J | 5650 hCoV-19/England/QEUH-9B8806/2020\|EPI_ISL_549620\|20 | ···················· | ···················· | ···················· | ···················· | ···················· | ···················· |
| J | 5651 hCoV-19/England/QEUH-9B8CDD/2020\|EPI_ISL_549621\|20 | ···················· | ···················· | ···················· | ···················· | ···················· | ···················· |
| J | 5652 hCoV-19/England/QEUH-9B92CB/2020\|EPI_ISL_549622\|20 | ···················· | ·A·················· | ···················· | ···················· | ···················· | ···A················ |
| J | 5653 hCoV-19/England/QEUH-9B8F98/2020\|EPI_ISL_549623\|20 | ···················· | ···················· | ···················· | ···················· | ···················· | ···············A···· |
| J | 5654 hCoV-19/England/QEUH-9B8C55/2020\|EPI_ISL_549624\|20 | ···················· | ···················· | ···················· | ···················· | ···················· | ···················· |
| J | 5655 hCoV-19/England/QEUH-9B9B48/2020\|EPI_ISL_549625\|20 | ···················· | ···················· | ···················· | ···················· | ···················· | ···················· |
| J | 5656 hCoV-19/England/QEUH-9B92DA/2020\|EPI_ISL_549626\|20 | ···················· | ···················· | ···················· | ···················· | ···················· | ···················· |
| J | 5657 hCoV-19/England/QEUH-9B887F/2020\|EPI_ISL_549627\|20 | ···················· | ···················· | ···················· | ···················· | ···················· | ···················· |
| J | 5658 hCoV-19/England/QEUH-9B9331/2020\|EPI_ISL_549628\|20 | ···················· | ······R············· | ···················· | ···················· | ···················· | ···················· |
| J | 5659 hCoV-19/England/QEUH-9B87FA/2020\|EPI_ISL_549629\|20 | ···················· | ·A·······A·········· | ···················· | ···················· | ···················· | ···················· |
| J | 5660 hCoV-19/England/QEUH-9B9D7F/2020\|EPI_ISL_549630\|20 | ···················· | ···················· | ···················· | ···················· | ···················· | ···················· |
| J | 5661 hCoV-19/England/QEUH-9B888E/2020\|EPI_ISL_549631\|20 | ···················· | ·A·················· | ···················· | ···················· | ···················· | ···A················ |
| J | 5662 hCoV-19/England/QEUH-9B8AC4/2020\|EPI_ISL_549632\|20 | ···················· | ···················· | ···················· | ···················· | ···················· | ···················· |
| J | 5663 hCoV-19/England/QEUH-9B8EF5/2020\|EPI_ISL_549633\|20 | ···················· | ···················· | ···················· | ···················· | ···················· | ···················· |
| J | 5664 hCoV-19/England/QEUH-9B99D5/2020\|EPI_ISL_549634\|20 | ···················· | ···················· | ···················· | ···················· | ···················· | ···················· |
| J | 5665 hCoV-19/England/QEUH-9B8E8C/2020\|EPI_ISL_549635\|20 | ···················· | ···················· | ···················· | ···················· | ···················· | ···················· |
| J | 5666 hCoV-19/England/QEUH-9B9B0C/2020\|EPI_ISL_549636\|20 | ···················· | ···················· | ···················· | ···················· | ···················· | ···················· |
| J | 5667 hCoV-19/England/QEUH-9B9B1B/2020\|EPI_ISL_549638\|20 | ···················· | ···················· | ···················· | ···················· | ···················· | ···················· |
| J | 5668 hCoV-19/England/QEUH-9B8323/2020\|EPI_ISL_549639\|20 | ···················· | ···················· | ···················· | ···················· | ···················· | ···················· |
| J | 5669 hCoV-19/England/QEUH-9B9AE1/2020\|EPI_ISL_549640\|20 | ···················· | ···················· | ···················· | ···················· | ···················· | ···················· |
| J | 5670 hCoV-19/England/QEUH-9B8A1F/2020\|EPI_ISL_549641\|20 | ···················· | ···················· | ···················· | ···················· | ···················· | ···················· |
| J | 5671 hCoV-19/England/QEUH-9B902B/2020\|EPI_ISL_549642\|20 | ···················· | ···················· | ···················· | ···················· | ···················· | ···················· |
| J | 5672 hCoV-19/England/QEUH-9B9CFA/2020\|EPI_ISL_549643\|20 | ···················· | ···················· | ···················· | ···················· | ···················· | ···················· |
| J | 5673 hCoV-19/England/QEUH-9B92F8/2020\|EPI_ISL_549644\|20 | ···················· | ···················· | ···················· | ···················· | ···················· | ···················· |

| | | | | | | | | |
|---|---|---|---|---|---|---|---|---|
| J | 5674 | hCoV-19/England/QEUH-9B9AF0/2020\|EPI_ISL_549645\|20 | ···················· | ···················· | ····················· | ···················· | ···················· | ···················· |
| J | 5675 | hCoV-19/England/QEUH-9B8BEF/2020\|EPI_ISL_549646\|20 | ···················· | ···················· | ····················· | ···················· | ···················· | ···················· |
| J | 5676 | hCoV-19/England/QEUH-9B8E22/2020\|EPI_ISL_549647\|20 | ···················· | ···················· | ····················· | ···················· | ···················· | ···················· |
| J | 5677 | hCoV-19/England/QEUH-9B92E9/2020\|EPI_ISL_549648\|20 | ···················· | ···················· | ····················· | ···················· | ···················· | ···················· |
| J | 5678 | hCoV-19/England/QEUH-9B90EF/2020\|EPI_ISL_549649\|20 | ···················· | ···················· | ····················· | ···················· | ···················· | ···················· |
| J | 5679 | hCoV-19/England/QEUH-9B8B1C/2020\|EPI_ISL_549651\|20 | ···················· | ···················· | ····················· | ···················· | ···················· | ···················· |
| J | 5680 | hCoV-19/England/QEUH-9B836F/2020\|EPI_ISL_549652\|20 | ···················· | ···················· | ····················· | ···················· | ···················· | ···················· |
| J | 5681 | hCoV-19/England/QEUH-9B903A/2020\|EPI_ISL_549653\|20 | ···················· | ···················· | ····················· | ···················· | ···················· | ···················· |
| J | 5682 | hCoV-19/England/QEUH-9B9164/2020\|EPI_ISL_549654\|20 | ···················· | ···················· | ····················· | ···················· | ···················· | ···················· |
| J | 5683 | hCoV-19/England/QEUH-9B9CDC/2020\|EPI_ISL_549655\|20 | ···················· | ···················· | ····················· | ···················· | ···················· | ···················· |
| J | 5684 | hCoV-19/England/QEUH-9B89B8/2020\|EPI_ISL_549656\|20 | ···················· | ···················· | ···T················· | ···················· | ···················· | ···················· |
| J | 5685 | hCoV-19/England/QEUH-9B8666/2020\|EPI_ISL_549657\|20 | ···················· | ···················· | ····················· | ···················· | ····T··············· | ···················· |
| J | 5686 | hCoV-19/England/QEUH-9B83D8/2020\|EPI_ISL_549658\|20 | ···················· | ···················· | ····················· | ···················· | ···················· | ···················· |
| J | 5687 | hCoV-19/England/QEUH-9B9C81/2020\|EPI_ISL_549659\|20 | ···················· | ···················· | ····················· | ···················· | ···················· | ···················· |
| J | 5688 | hCoV-19/England/MILK-99DA10/2020\|EPI_ISL_550600\|20 | ···················· | ···················· | ····················· | ···················· | ···················· | ···················· |
| J | 5689 | hCoV-19/England/MILK-99DBC2/2020\|EPI_ISL_550601\|20 | ···················· | ·A·················· | ····················· | ···················· | ···················· | ···················· |
| J | 5690 | hCoV-19/England/MILK-99D209/2020\|EPI_ISL_550602\|20 | ···················· | ···················· | ····················· | ···················· | ···················· | ···················· |
| J | 5691 | hCoV-19/England/MILK-99C729/2020\|EPI_ISL_550603\|20 | ···················· | ···················· | ····················· | ···················· | ···················· | ···················· |
| J | 5692 | hCoV-19/England/MILK-99C89F/2020\|EPI_ISL_550604\|20 | ···················· | ···················· | ····················· | ···················· | ···················· | ···················· |
| J | 5693 | hCoV-19/England/MILK-99DB59/2020\|EPI_ISL_550605\|20 | ···················· | ···················· | ····················· | ···················· | ···················· | ···················· |
| J | 5694 | hCoV-19/England/MILK-99D324/2020\|EPI_ISL_550606\|20 | ···················· | ·A·················· | ····················· | ···················· | ···················· | ···A················ |
| J | 5695 | hCoV-19/England/MILK-99C97E/2020\|EPI_ISL_550607\|20 | ···················· | ···················· | ····················· | ···················· | ···················· | ···················· |
| J | 5696 | hCoV-19/England/MILK-99D236/2020\|EPI_ISL_550608\|20 | ···················· | ···················· | ····················· | ···················· | ···················· | ···················· |
| J | 5697 | hCoV-19/England/MILK-99E411/2020\|EPI_ISL_550609\|20 | ···················· | ···················· | ····················· | ···················· | ···················· | ···················· |
| J | 5698 | hCoV-19/England/MILK-99E45D/2020\|EPI_ISL_550610\|20 | ···················· | ···················· | ····················· | ···················· | ···················· | ···················· |
| J | 5699 | hCoV-19/England/MILK-99E192/2020\|EPI_ISL_550611\|20 | ···················· | ···················· | ····················· | ···················· | ···················· | ···················· |
| J | 5700 | hCoV-19/England/MILK-99DB0E/2020\|EPI_ISL_550612\|20 | ···················· | ···················· | ····················· | ···················· | ···················· | ···················· |
| J | 5701 | hCoV-19/England/MILK-99DFF3/2020\|EPI_ISL_550613\|20 | ···················· | ···················· | ····················· | ···················· | ···················· | ···················· |
| J | 5702 | hCoV-19/Wales/MILK-99C53E/2020\|EPI_ISL_550614\|2020 | ···················· | ···················· | ····················· | ···················· | ···················· | ···················· |
| J | 5703 | hCoV-19/England/MILK-99E3D8/2020\|EPI_ISL_550615\|20 | ···················· | ···················· | ····················· | ···················· | ···················· | ···················· |
| J | 5704 | hCoV-19/England/MILK-99C941/2020\|EPI_ISL_550616\|20 | ···················· | ···················· | ····················· | ···················· | ···················· | ···················· |
| J | 5705 | hCoV-19/England/MILK-99E44E/2020\|EPI_ISL_550617\|20 | ···················· | ···················· | ····················· | ···················· | ···················· | ···················· |
| J | 5706 | hCoV-19/England/MILK-99CB4B/2020\|EPI_ISL_550618\|20 | ···················· | ···················· | ····················· | ···················· | ···················· | ···················· |
| J | 5707 | hCoV-19/England/MILK-99D99B/2020\|EPI_ISL_550619\|20 | ···················· | ·A·················· | ····················· | ···················· | ···················· | ···A················ |
| J | 5708 | hCoV-19/England/MILK-99DB86/2020\|EPI_ISL_550620\|20 | ···················· | ···················· | ····················· | ···················· | ···················· | ···················· |
| J | 5709 | hCoV-19/England/MILK-99D9F5/2020\|EPI_ISL_550621\|20 | ···················· | ···················· | ····················· | ···················· | ···················· | ···················· |
| J | 5710 | hCoV-19/England/MILK-99E165/2020\|EPI_ISL_550622\|20 | ···················· | ···················· | ····················· | ···················· | ···················· | ···················· |
| J | 5711 | hCoV-19/England/MILK-99DDF9/2020\|EPI_ISL_550625\|20 | ···················· | ···················· | ····················· | ···················· | ···················· | ···················· |
| J | 5712 | hCoV-19/England/MILK-99D245/2020\|EPI_ISL_550626\|20 | ···················· | ···················· | ····················· | ···················· | ···················· | ···················· |
| J | 5713 | hCoV-19/England/MILK-99DB1D/2020\|EPI_ISL_550627\|20 | ···················· | ···················· | ····················· | ···················· | ···················· | ···················· |
| J | 5714 | hCoV-19/England/MILK-99E47B/2020\|EPI_ISL_550628\|20 | ···················· | ····A··············· | ····················· | ···················· | ···················· | ···················· |
| J | 5715 | hCoV-19/England/MILK-99E5D2/2020\|EPI_ISL_550629\|20 | ···················· | ···················· | ····················· | ···················· | ···················· | ···················· |
| J | 5716 | hCoV-19/England/MILK-99DE50/2020\|EPI_ISL_550630\|20 | ···················· | ···················· | ····················· | ···················· | ···················· | ···················· |
| J | 5717 | hCoV-19/England/MILK-99CA20/2020\|EPI_ISL_550631\|20 | ···················· | ···················· | ····················· | ···················· | ···················· | ···················· |
| J | 5718 | hCoV-19/England/MILK-99E280/2020\|EPI_ISL_550632\|20 | ···················· | ···················· | ····················· | ···················· | ···················· | ···················· |
| J | 5719 | hCoV-19/England/MILK-99C4B9/2020\|EPI_ISL_550633\|20 | ········T··········· | ···················· | ····················· | ···················· | ···················· | ···················· |
| J | 5720 | hCoV-19/England/MILK-99E068/2020\|EPI_ISL_550634\|20 | ···················· | ···················· | ····················· | ···················· | ···················· | ···················· |
| J | 5721 | hCoV-19/England/MILK-99C6E0/2020\|EPI_ISL_550635\|20 | ···················· | ···················· | ····················· | ···················· | ······T············· | ···················· |
| J | 5722 | hCoV-19/England/MILK-99DC92/2020\|EPI_ISL_550636\|20 | ···················· | ···················· | ····················· | ···················· | ···················· | ···················· |
| J | 5723 | hCoV-19/England/MILK-99E402/2020\|EPI_ISL_550637\|20 | ···················· | ···················· | ····················· | ···················· | ···················· | ···················· |
| J | 5724 | hCoV-19/Wales/MILK-99C4AA/2020\|EPI_ISL_550638\|2020 | ···················· | ···················· | ····················· | ···················· | ···················· | ···················· |
| J | 5725 | hCoV-19/England/CAMC-990F2D/2020\|EPI_ISL_551900\|20 | ···················· | ···················· | ····················· | ···················· | ···················· | ···················· |
| J | 5726 | hCoV-19/England/CAMC-990A1D/2020\|EPI_ISL_551904\|20 | ···················· | ···················· | ····················· | ···················· | ···················· | ···················· |
| J | 5727 | hCoV-19/England/CAMC-99094D/2020\|EPI_ISL_551910\|20 | ···················· | ···················· | ····················· | ···················· | ···················· | ···················· |
| J | 5728 | hCoV-19/England/CAMC-99100B/2020\|EPI_ISL_551911\|20 | ···················· | ···················· | ····················· | ···················· | ···················· | ···················· |
| J | 5729 | hCoV-19/England/CAMC-990D7E/2020\|EPI_ISL_551912\|20 | ···················· | ·A·················· | ····················· | ···················· | ···················· | ···················· |
| J | 5730 | hCoV-19/England/CAMC-990D8D/2020\|EPI_ISL_551913\|20 | ···················· | ···················· | ····················· | ···················· | ···················· | ···················· |
| J | 5731 | hCoV-19/England/CAMC-9908E6/2020\|EPI_ISL_551914\|20 | ···················· | ···················· | ····················· | ···················· | ···················· | ···················· |
| J | 5732 | hCoV-19/England/CAMC-990EE4/2020\|EPI_ISL_551915\|20 | ···················· | ···················· | ····················· | ···················· | ···················· | ···················· |
| J | 5733 | hCoV-19/England/CAMC-990E99/2020\|EPI_ISL_551916\|20 | ···················· | ···················· | ····················· | ···················· | ······T············· | ···················· |
| J | 5734 | hCoV-19/England/CAMC-9910ED/2020\|EPI_ISL_551917\|20 | ···················· | ···················· | ····················· | ···················· | ···················· | ···················· |
| J | 5735 | hCoV-19/England/CAMC-990BFC/2020\|EPI_ISL_551918\|20 | ···················· | ···················· | ····················· | ···················· | ···················· | ···················· |
| J | 5736 | hCoV-19/England/CAMC-9909E3/2020\|EPI_ISL_551919\|20 | ···················· | ···················· | ····················· | ···················· | ···················· | ···················· |
| J | 5737 | hCoV-19/England/MILK-99341C/2020\|EPI_ISL_551923\|20 | ···················· | ···················· | ····················· | ···················· | ···················· | ···················· |
| J | 5738 | hCoV-19/England/MILK-993379/2020\|EPI_ISL_551924\|20 | ···················· | ···················· | ····················· | ···················· | ···················· | ···················· |
| J | 5739 | hCoV-19/England/CAMC-991706/2020\|EPI_ISL_551925\|20 | ···················· | ···················· | ····················· | ···················· | ···················· | ···················· |
| J | 5740 | hCoV-19/England/MILK-992802/2020\|EPI_ISL_551926\|20 | ···················· | ···················· | ····················· | ···················· | ···················· | ···················· |
| J | 5741 | hCoV-19/Wales/MILK-992C15/2020\|EPI_ISL_551927\|2020 | ···················· | ···················· | ····················· | ···················· | ···················· | ···················· |
| J | 5742 | hCoV-19/England/MILK-993661/2020\|EPI_ISL_551928\|20 | ···················· | ···················· | ····················· | ···················· | ···················· | ···················· |
| J | 5743 | hCoV-19/England/MILK-992714/2020\|EPI_ISL_551929\|20 | ···················· | ···················· | ····················· | ···················· | ···················· | ···················· |
| J | 5744 | hCoV-19/England/MILK-993449/2020\|EPI_ISL_551930\|20 | ···················· | ···················· | ····················· | ···················· | ···················· | ···················· |
| J | 5745 | hCoV-19/England/MILK-99292D/2020\|EPI_ISL_551931\|20 | ···················· | ···················· | ····················· | ···················· | ···················· | ···················· |
| J | 5746 | hCoV-19/England/CAMC-99152A/2020\|EPI_ISL_551932\|20 | ···················· | ···················· | ····················· | ···················· | ···················· | ···················· |
| J | 5747 | hCoV-19/England/MILK-9936AD/2020\|EPI_ISL_551934\|20 | ···················· | ···················· | ····················· | ···················· | ···················· | ···················· |
| J | 5748 | hCoV-19/England/MILK-992DF4/2020\|EPI_ISL_551935\|20 | ···················· | ···················· | ····················· | ···················· | ···················· | ···················· |
| J | 5749 | hCoV-19/England/MILK-9935A0/2020\|EPI_ISL_551936\|20 | ···················· | ···················· | ····················· | ···················· | ···················· | ···················· |
| J | 5750 | hCoV-19/Wales/MILK-993142/2020\|EPI_ISL_551938\|2020 | ···················· | ···················· | ····················· | ···················· | ···················· | ···················· |
| J | 5751 | hCoV-19/England/MILK-992C9D/2020\|EPI_ISL_551939\|20 | ···················· | ···················· | ····················· | ···················· | ···················· | ···················· |
| J | 5752 | hCoV-19/England/MILK-992723/2020\|EPI_ISL_551940\|20 | ···················· | ···················· | ····················· | ···················· | ···················· | ···················· |
| J | 5753 | hCoV-19/England/MILK-992A1B/2020\|EPI_ISL_551942\|20 | ···················· | ···················· | ····················· | ···················· | ···················· | ···················· |

| | | | | | | | | |
|---|---|---|---|---|---|---|---|---|
| J | 5754 | hCoV-19/England/MILK-991F0E/2020\|EPI_ISL_551943\|20 | ···················· | ·A·················· | ···················· | ···················· | ···················· | ···················· |
| J | 5755 | hCoV-19/England/MILK-9932E5/2020\|EPI_ISL_551944\|20 | ···················· | ···················· | ···················· | ···················· | ······T············· | ···················· |
| J | 5756 | hCoV-19/England/MILK-9937C8/2020\|EPI_ISL_551945\|20 | ···················· | ···················· | ···················· | ···················· | ···················· | ···················· |
| J | 5757 | hCoV-19/England/MILK-9927BA/2020\|EPI_ISL_551946\|20 | ···················· | ···················· | ···················· | ···················· | ···················· | ···················· |
| J | 5758 | hCoV-19/England/MILK-991F1D/2020\|EPI_ISL_551947\|20 | ···················· | ···················· | ···················· | ···················· | ···················· | ···················· |
| J | 5759 | hCoV-19/England/MILK-991E4D/2020\|EPI_ISL_551948\|20 | ···················· | ···················· | ···················· | ···················· | ···················· | ···················· |
| J | 5760 | hCoV-19/Wales/MILK-992CF7/2020\|EPI_ISL_551949\|2020 | ···················· | ···················· | ···················· | ···················· | ···················· | ···················· |
| J | 5761 | hCoV-19/England/MILK-9933F1/2020\|EPI_ISL_551950\|20 | ···················· | ···················· | ···················· | ···················· | ···················· | ···················· |
| J | 5762 | hCoV-19/England/CAMC-99143C/2020\|EPI_ISL_551951\|20 | ···················· | ···················· | ···················· | ···················· | ···················· | ···················· |
| J | 5763 | hCoV-19/England/MILK-9931E8/2020\|EPI_ISL_551952\|20 | ···················· | ···················· | ···················· | ···················· | ···················· | ···················· |
| J | 5764 | hCoV-19/England/CAMC-991618/2020\|EPI_ISL_551953\|20 | ···················· | ···················· | ···················· | ···················· | ···················· | ···················· |
| J | 5765 | hCoV-19/England/MILK-992ED3/2020\|EPI_ISL_551954\|20 | ···················· | ···················· | ···················· | ···················· | ···················· | ···················· |
| J | 5766 | hCoV-19/England/MILK-992C06/2020\|EPI_ISL_551955\|20 | ···················· | ···················· | ···················· | ···················· | ···················· | ···················· |
| J | 5767 | hCoV-19/England/MILK-9937AA/2020\|EPI_ISL_551956\|20 | ···················· | ···················· | ···················· | ···················· | ···················· | ···················· |
| J | 5768 | hCoV-19/England/MILK-992D7C/2020\|EPI_ISL_551957\|20 | ···················· | ···················· | ···················· | ···················· | ···················· | ···················· |
| J | 5769 | hCoV-19/England/MILK-992B18/2020\|EPI_ISL_551958\|20 | ···················· | ···················· | ···················· | ···················· | ···················· | ···················· |
| J | 5770 | hCoV-19/England/MILK-9932C7/2020\|EPI_ISL_551959\|20 | ···················· | ·A·················· | ···················· | ···················· | ···················· | ··A················· |
| J | 5771 | hCoV-19/England/MILK-992A39/2020\|EPI_ISL_551960\|20 | ···················· | ···················· | ···················· | ···················· | ···················· | ···················· |
| J | 5772 | hCoV-19/England/MILK-993300/2020\|EPI_ISL_551962\|20 | ···················· | ···················· | ···················· | ···················· | ···················· | ···················· |
| J | 5773 | hCoV-19/England/MILK-992F2B/2020\|EPI_ISL_551963\|20 | ···················· | ···················· | ···················· | ···················· | ···················· | ···················· |
| J | 5774 | hCoV-19/England/MILK-992978/2020\|EPI_ISL_551965\|20 | ···················· | ·A·······A·········· | ···················· | ···················· | ···················· | ···················· |
| J | 5775 | hCoV-19/England/MILK-992D30/2020\|EPI_ISL_551966\|20 | ···················· | ···················· | ···················· | ···················· | ···················· | ···················· |
| J | 5776 | hCoV-19/England/CAMC-991548/2020\|EPI_ISL_551967\|20 | ···················· | ···················· | ···················· | ···················· | ···················· | ···················· |
| J | 5777 | hCoV-19/England/MILK-99327C/2020\|EPI_ISL_551968\|20 | ···················· | ···················· | ···················· | ···················· | ···················· | ···················· |
| J | 5778 | hCoV-19/England/MILK-99375F/2020\|EPI_ISL_551969\|20 | ···················· | ···················· | ···················· | ···················· | ···················· | ···················· |
| J | 5779 | hCoV-19/England/MILK-993713/2020\|EPI_ISL_551970\|20 | ···················· | ···················· | ···················· | ···················· | ···················· | ···················· |
| J | 5780 | hCoV-19/England/MILK-957371/2020\|EPI_ISL_554100\|20 | ···················· | ···················· | ···················· | ···················· | ···················· | ···················· |
| J | 5781 | hCoV-19/England/MILK-9573EA/2020\|EPI_ISL_554101\|20 | ···················· | ···················· | ···················· | ···················· | ···················· | ···················· |
| J | 5782 | hCoV-19/England/ALDP-952859/2020\|EPI_ISL_554102\|20 | ···················· | ···················· | ···················· | ···················· | ···················· | ···················· |
| J | 5783 | hCoV-19/England/ALDP-956309/2020\|EPI_ISL_554104\|20 | ···················· | ···················· | ···················· | ···················· | ···················· | ···················· |
| J | 5784 | hCoV-19/England/ALDP-9563CD/2020\|EPI_ISL_554105\|20 | ···················· | ···················· | ···················· | ···················· | ···················· | ···················· |
| J | 5785 | hCoV-19/England/ALDP-95277A/2020\|EPI_ISL_554106\|20 | ···················· | ···················· | ···················· | ···················· | ···················· | ···················· |
| J | 5786 | hCoV-19/Wales/ALDP-955EDC/2020\|EPI_ISL_554107\|2020 | ········Y··········· | ···················· | ···················· | ···················· | ···················· | ···················· |
| J | 5787 | hCoV-19/England/ALDP-9528B3/2020\|EPI_ISL_554108\|20 | ···················· | ···················· | ···················· | ···················· | ···················· | ···················· |
| J | 5788 | hCoV-19/England/ALDP-952ADB/2020\|EPI_ISL_554109\|20 | ···················· | ···················· | ···················· | ···················· | ···················· | ···················· |
| J | 5789 | hCoV-19/England/ALDP-955FAC/2020\|EPI_ISL_554110\|20 | ···················· | ···················· | ···················· | ···················· | ···················· | ···················· |
| J | 5790 | hCoV-19/England/ALDP-95603F/2020\|EPI_ISL_554111\|20 | ···················· | ···················· | ···················· | ···················· | ···················· | ···················· |
| J | 5791 | hCoV-19/England/ALDP-9563DC/2020\|EPI_ISL_554112\|20 | ···················· | ···················· | ···················· | ···················· | ···················· | ···················· |
| J | 5792 | hCoV-19/England/ALDP-955F06/2020\|EPI_ISL_554113\|20 | ···················· | ···················· | ···················· | ···················· | ···················· | ···················· |
| J | 5793 | hCoV-19/England/ALDP-9527C5/2020\|EPI_ISL_554114\|20 | ···················· | ···················· | ···················· | ···················· | ···················· | ···················· |
| J | 5794 | hCoV-19/England/ALDP-9564D9/2020\|EPI_ISL_554116\|20 | ···················· | ···················· | ···················· | ···················· | ···················· | ···················· |
| J | 5795 | hCoV-19/England/ALDP-95280E/2020\|EPI_ISL_554117\|20 | ···················· | ···················· | ···················· | ···················· | ···················· | ···················· |
| J | 5796 | hCoV-19/England/MILK-956CE0/2020\|EPI_ISL_554118\|20 | ···················· | ···················· | ···················· | ···················· | ···················· | ···················· |
| J | 5797 | hCoV-19/England/MILK-957292/2020\|EPI_ISL_554121\|20 | ···················· | ···················· | ···················· | ···················· | ···················· | ···················· |
| J | 5798 | hCoV-19/England/ALDP-95177B/2020\|EPI_ISL_554122\|20 | ···················· | ···················· | ···················· | ···················· | ···················· | ···················· |
| J | 5799 | hCoV-19/England/MILK-9572CF/2020\|EPI_ISL_554123\|20 | ···················· | ···················· | ···················· | ···················· | ···················· | ···················· |
| J | 5800 | hCoV-19/England/MILK-9567C1/2020\|EPI_ISL_554124\|20 | ···················· | ···················· | ···················· | ···················· | ···················· | ···················· |
| J | 5801 | hCoV-19/England/MILK-957B79/2020\|EPI_ISL_554125\|20 | ···················· | ···················· | ···················· | ···················· | ···················· | ···················· |
| J | 5802 | hCoV-19/England/MILK-956C2C/2020\|EPI_ISL_554134\|20 | ···················· | ···················· | ···················· | ···················· | ···················· | ···················· |
| J | 5803 | hCoV-19/England/ALDP-951061/2020\|EPI_ISL_554136\|20 | ···················· | ···················· | ···················· | ···················· | ···················· | ···················· |
| J | 5804 | hCoV-19/England/MILK-956E53/2020\|EPI_ISL_554137\|20 | ···················· | ···················· | ···················· | ···················· | ···················· | ···················· |
| J | 5805 | hCoV-19/England/ALDP-951571/2020\|EPI_ISL_554190\|20 | ···················· | ···················· | ···················· | ···················· | ···················· | ···················· |
| J | 5806 | hCoV-19/England/MILK-956A31/2020\|EPI_ISL_554191\|20 | ···················· | ···················· | ···················· | ···················· | ···················· | ···················· |
| J | 5807 | hCoV-19/England/ALDP-950E95/2020\|EPI_ISL_554192\|20 | ···················· | ···················· | ···················· | ···················· | ···················· | ···················· |
| J | 5808 | hCoV-19/England/ALDP-950FCF/2020\|EPI_ISL_554194\|20 | ···················· | ···················· | ···················· | ···················· | ···················· | ···················· |
| J | 5809 | hCoV-19/England/ALDP-950F92/2020\|EPI_ISL_554196\|20 | ···················· | ·A·················· | ···················· | ···················· | ···················· | ···················· |
| J | 5810 | hCoV-19/England/ALDP-950F83/2020\|EPI_ISL_554197\|20 | ···················· | ·A·················· | ···················· | ···················· | ···················· | ···················· |
| J | 5811 | hCoV-19/England/ALDP-95118C/2020\|EPI_ISL_554198\|20 | ···················· | ···················· | ···················· | ···················· | ···················· | ···················· |
| J | 5812 | hCoV-19/England/ALDP-9513C2/2020\|EPI_ISL_554199\|20 | ···················· | ···················· | ···················· | ···················· | ···················· | ···················· |
| J | 5813 | hCoV-19/England/ALDP-955A8D/2020\|EPI_ISL_555414\|20 | ···················· | ···················· | ···················· | ···················· | ···················· | ···················· |
| J | 5814 | hCoV-19/England/ALDP-94E51D/2020\|EPI_ISL_555415\|20 | ···················· | ···················· | ···················· | ···················· | ···················· | ···················· |
| J | 5815 | hCoV-19/England/ALDP-95472D/2020\|EPI_ISL_555416\|20 | ···················· | ···················· | ···················· | ···················· | ···················· | ···················· |
| J | 5816 | hCoV-19/England/ALDP-955BF3/2020\|EPI_ISL_555417\|20 | ···················· | ···················· | ···················· | ···················· | ···················· | ···················· |
| J | 5817 | hCoV-19/England/ALDP-9519FD/2020\|EPI_ISL_555418\|20 | ···················· | ···················· | ···················· | ···················· | ···················· | ···················· |
| J | 5818 | hCoV-19/England/ALDP-9548DF/2020\|EPI_ISL_555420\|20 | ···················· | ···················· | ···················· | ···················· | ···················· | ···················· |
| J | 5819 | hCoV-19/England/ALDP-955A41/2020\|EPI_ISL_555421\|20 | ···················· | ···················· | ···················· | ···················· | ···················· | ···················· |
| J | 5820 | hCoV-19/England/ALDP-952B41/2020\|EPI_ISL_555423\|20 | ···················· | ···················· | ···················· | ···················· | ···················· | ···················· |
| J | 5821 | hCoV-19/England/ALDP-954893/2020\|EPI_ISL_555424\|20 | ···················· | ···················· | ···················· | ···················· | ···················· | ···················· |
| J | 5822 | hCoV-19/England/ALDP-951A27/2020\|EPI_ISL_555425\|20 | ···················· | ···················· | ···················· | ···················· | ···················· | ···················· |
| J | 5823 | hCoV-19/England/ALDP-9547F0/2020\|EPI_ISL_555426\|20 | ···················· | ···················· | ···················· | ···················· | ···················· | ···················· |
| J | 5824 | hCoV-19/England/ALDP-954884/2020\|EPI_ISL_555427\|20 | ···················· | ···················· | ···················· | ···················· | ···················· | ···················· |
| J | 5825 | hCoV-19/England/ALDP-94E128/2020\|EPI_ISL_555428\|20 | ···················· | ···················· | ···················· | ···················· | ···················· | ···················· |
| J | 5826 | hCoV-19/England/ALDP-954523/2020\|EPI_ISL_555429\|20 | ···················· | ···················· | ···················· | ···················· | ···················· | ···················· |
| J | 5827 | hCoV-19/England/ALDP-954778/2020\|EPI_ISL_555430\|20 | ···················· | ···················· | ···················· | ···················· | ···················· | ···················· |
| J | 5828 | hCoV-19/England/ALDP-952B7E/2020\|EPI_ISL_555431\|20 | ···················· | ···················· | ···················· | ···················· | ···················· | ···················· |
| J | 5829 | hCoV-19/England/ALDP-955A7E/2020\|EPI_ISL_555435\|20 | ···················· | ···················· | ···················· | ···················· | ···················· | ···················· |
| J | 5830 | hCoV-19/England/ALDP-94E665/2020\|EPI_ISL_555436\|20 | ···················· | ···················· | ···················· | ···················· | ···················· | ···················· |
| J | 5831 | hCoV-19/Wales/ALDP-94E6B0/2020\|EPI_ISL_555438\|2020 | ···················· | ···················· | ···················· | ···················· | ···················· | ···················· |
| J | 5832 | hCoV-19/England/ALDP-955A23/2020\|EPI_ISL_555439\|20 | ···················· | ···················· | ···················· | ···················· | ···················· | ···················· |
| J | 5833 | hCoV-19/England/ALDP-94E744/2020\|EPI_ISL_555441\|20 | ···················· | ···················· | ···················· | ···················· | ···················· | ···················· |

| | | | | | | | | |
|---|---|---|---|---|---|---|---|---|
| J | 5834 | hCoV-19/England/ALDP-954514/2020\|EPI_ISL_555442\|20 | ···················· | ···················· | ····················· | ···················· | ···················· | ···················· |
| J | 5835 | hCoV-19/England/ALDP-94E753/2020\|EPI_ISL_555443\|20 | ···················· | ···················· | ····················· | ···················· | ···················· | ···················· |
| J | 5836 | hCoV-19/England/ALDP-951A45/2020\|EPI_ISL_555444\|20 | ···················· | ···················· | ····················· | ···················· | ···················· | ···················· |
| J | 5837 | hCoV-19/England/ALDP-95599F/2020\|EPI_ISL_555445\|20 | ···················· | ···················· | ····················· | ···················· | ···················· | ···················· |
| J | 5838 | hCoV-19/England/ALDP-955C4B/2020\|EPI_ISL_555447\|20 | ···················· | ···················· | ····················· | ···················· | ···················· | ···················· |
| J | 5839 | hCoV-19/England/ALDP-95457E/2020\|EPI_ISL_555448\|20 | ···················· | ···················· | ····················· | ···················· | ···················· | ···················· |
| J | 5840 | hCoV-19/England/ALDP-94E3AA/2020\|EPI_ISL_555449\|20 | ···················· | ···················· | ····················· | ···················· | ···················· | ···················· |
| J | 5841 | hCoV-19/England/ALDP-94E42F/2020\|EPI_ISL_555450\|20 | ···················· | ···················· | ····················· | ···················· | ···················· | ···················· |
| J | 5842 | hCoV-19/England/ALDP-951CF4/2020\|EPI_ISL_555452\|20 | ···················· | ···················· | ····················· | ···················· | ···················· | ···················· |
| J | 5843 | hCoV-19/England/ALDP-954550/2020\|EPI_ISL_555453\|20 | ···················· | ···················· | ····················· | ···················· | ···················· | ···················· |
| J | 5844 | hCoV-19/England/ALDP-9518D2/2020\|EPI_ISL_555454\|20 | ···················· | ···················· | ····················· | ···················· | ···················· | ···················· |
| J | 5845 | hCoV-19/England/ALDP-951887/2020\|EPI_ISL_555456\|20 | ···················· | ···················· | ····················· | ···················· | ···················· | ···················· |
| J | 5846 | hCoV-19/England/ALDP-94E53B/2020\|EPI_ISL_555457\|20 | ···················· | ···················· | ····················· | ···················· | ···················· | ···················· |
| J | 5847 | hCoV-19/England/ALDP-951AFA/2020\|EPI_ISL_555468\|20 | ···················· | ···················· | ····················· | ···················· | ···················· | ···················· |
| J | 5848 | hCoV-19/England/CAMC-945B32/2020\|EPI_ISL_556745\|20 | ···················· | ···················· | ····················· | ···················· | ···················· | ···················· |
| J | 5849 | hCoV-19/England/CAMC-94A748/2020\|EPI_ISL_556747\|20 | ···················· | ···················· | ····················· | ···················· | ···················· | ···················· |
| J | 5850 | hCoV-19/England/CAMC-94B0A6/2020\|EPI_ISL_556748\|20 | ···················· | ···················· | ····················· | ···················· | ···················· | ···················· |
| J | 5851 | hCoV-19/England/CAMC-9457B6/2020\|EPI_ISL_556749\|20 | ···················· | ···················· | ····················· | ···················· | ···················· | ···················· |
| J | 5852 | hCoV-19/England/CAMC-949C68/2020\|EPI_ISL_556750\|20 | ···················· | ···················· | ····················· | ···················· | ···················· | ···················· |
| J | 5853 | hCoV-19/England/CAMC-94A784/2020\|EPI_ISL_556752\|20 | ···················· | ·A·················· | ····················· | ···················· | ···················· | ···················· |
| J | 5854 | hCoV-19/England/CAMC-949F7E/2020\|EPI_ISL_556753\|20 | ···················· | ···················· | ····················· | ···················· | ···················· | ···················· |
| J | 5855 | hCoV-19/England/CAMC-948A6F/2020\|EPI_ISL_556754\|20 | ···················· | ······A············· | ····················· | ···················· | ···················· | ···················· |
| J | 5856 | hCoV-19/England/CAMC-94B57A/2020\|EPI_ISL_556755\|20 | ···················· | ···················· | ····················· | ···················· | ···················· | ···················· |
| J | 5857 | hCoV-19/England/CAMC-94BC93/2020\|EPI_ISL_556756\|20 | ···················· | ···················· | ····················· | ···················· | ···················· | ···················· |
| J | 5858 | hCoV-19/England/CAMC-9491C3/2020\|EPI_ISL_556757\|20 | ···················· | ·A·················· | ····················· | ···················· | ···················· | ···················· |
| J | 5859 | hCoV-19/England/CAMC-94BBD2/2020\|EPI_ISL_556758\|20 | ···················· | ···················· | ····················· | ···················· | ···················· | ···················· |
| J | 5860 | hCoV-19/England/CAMC-94874A/2020\|EPI_ISL_556760\|20 | ···················· | ·A·················· | ····················· | ···················· | ···················· | ···················· |
| J | 5861 | hCoV-19/England/CAMC-946D2C/2020\|EPI_ISL_556762\|20 | ···················· | ···················· | ····················· | ···················· | ···················· | ···················· |
| J | 5862 | hCoV-19/England/CAMC-945B23/2020\|EPI_ISL_556763\|20 | ···················· | ···················· | ····················· | ···················· | ···················· | ···················· |
| J | 5863 | hCoV-19/England/CAMC-948B5D/2020\|EPI_ISL_556764\|20 | ···················· | ···················· | ···········T········· | ···················· | ···················· | ···················· |
| J | 5864 | hCoV-19/England/CAMC-949196/2020\|EPI_ISL_556765\|20 | ···················· | ···················· | ····················· | ···················· | ···················· | ···················· |
| J | 5865 | hCoV-19/England/CAMC-9491A5/2020\|EPI_ISL_556766\|20 | ···················· | ···················· | ····················· | ···················· | ···················· | ···················· |
| J | 5866 | hCoV-19/England/CAMC-949327/2020\|EPI_ISL_556767\|20 | ···················· | ·A·················· | ····················· | ···················· | ···················· | ···A················ |
| J | 5867 | hCoV-19/England/CAMC-946B31/2020\|EPI_ISL_556768\|20 | ···················· | ···················· | ····················· | ···················· | ···················· | ···················· |
| J | 5868 | hCoV-19/England/CAMC-94BC84/2020\|EPI_ISL_556769\|20 | ···················· | ···················· | ····················· | ···················· | ···················· | ···················· |
| J | 5869 | hCoV-19/England/CAMC-94971C/2020\|EPI_ISL_556771\|20 | ···················· | ·A·················· | ····················· | ···················· | ···················· | ···················· |
| J | 5870 | hCoV-19/England/CAMC-94BBF0/2020\|EPI_ISL_556772\|20 | ···················· | ···················· | ····················· | ···················· | ···················· | ···················· |
| J | 5871 | hCoV-19/England/CAMC-94B923/2020\|EPI_ISL_556773\|20 | ···················· | ···················· | ····················· | ···················· | ···················· | ···················· |
| J | 5872 | hCoV-19/England/CAMC-945701/2020\|EPI_ISL_556774\|20 | ···················· | ···················· | ····················· | ···················· | ···················· | ···················· |
| J | 5873 | hCoV-19/England/CAMC-946DB3/2020\|EPI_ISL_556775\|20 | ···················· | ···················· | ····················· | ···················· | ···················· | ···················· |
| J | 5874 | hCoV-19/England/CAMC-94BC39/2020\|EPI_ISL_556776\|20 | ···················· | ·A·················· | ····················· | ···················· | ···················· | ···················· |
| J | 5875 | hCoV-19/England/CAMC-94C02D/2020\|EPI_ISL_556777\|20 | ···················· | ·A·················· | ····················· | ···················· | ···················· | ···················· |
| J | 5876 | hCoV-19/England/CAMC-9458B3/2020\|EPI_ISL_556778\|20 | ···················· | ···················· | ····················· | ···················· | ···················· | ···················· |
| J | 5877 | hCoV-19/England/CAMC-94C01E/2020\|EPI_ISL_556779\|20 | ···················· | ···················· | ····················· | ···················· | ···················· | ···················· |
| J | 5878 | hCoV-19/England/CAMC-94BC57/2020\|EPI_ISL_556780\|20 | ···················· | ···················· | ····················· | ···················· | ···················· | ···················· |
| J | 5879 | hCoV-19/England/CAMC-949BC5/2020\|EPI_ISL_556781\|20 | ···················· | ···················· | ····················· | ···················· | ···················· | ···················· |
| J | 5880 | hCoV-19/England/CAMC-9493FA/2020\|EPI_ISL_556782\|20 | ···················· | ···················· | ····················· | ···················· | ···················· | ···················· |
| J | 5881 | hCoV-19/England/CAMC-94A502/2020\|EPI_ISL_556783\|20 | ···················· | ···················· | ····················· | ···················· | ···················· | ···················· |
| J | 5882 | hCoV-19/Wales/CAMC-94AFE6/2020\|EPI_ISL_556784\|2020 | ···················· | ···················· | ····················· | ···················· | ···················· | ···················· |
| J | 5883 | hCoV-19/England/CAMC-94A960/2020\|EPI_ISL_556785\|20 | ···················· | ···················· | ····················· | ···················· | ···················· | ···················· |
| J | 5884 | hCoV-19/England/CAMC-9454DD/2020\|EPI_ISL_556786\|20 | ···················· | ···················· | ····················· | ···················· | ···················· | ···················· |
| J | 5885 | hCoV-19/England/CAMC-94531C/2020\|EPI_ISL_556788\|20 | ···················· | ···················· | ····················· | ···················· | ···················· | ···················· |
| J | 5886 | hCoV-19/England/CAMC-946E65/2020\|EPI_ISL_556797\|20 | ···················· | ···················· | ····················· | ···················· | ···················· | ···················· |
| J | 5887 | hCoV-19/England/CAMC-949B89/2020\|EPI_ISL_556798\|20 | ···················· | ···················· | ····················· | ···················· | ···················· | ···················· |
| J | 5888 | hCoV-19/England/CAMC-94BB1E/2020\|EPI_ISL_556799\|20 | ···················· | ···················· | ···········T········· | ···················· | ···················· | ···················· |
| J | 5889 | hCoV-19/England/QEUH-9B8A2E/2020\|EPI_ISL_549660\|20 | ···················· | ···················· | ····················· | ···················· | ···················· | ···················· |
| J | 5890 | hCoV-19/England/QEUH-9B8851/2020\|EPI_ISL_549661\|20 | ···················· | ···················· | ····················· | ···················· | ···················· | ···················· |
| J | 5891 | hCoV-19/England/QEUH-9B8F7A/2020\|EPI_ISL_549662\|20 | ···················· | ···················· | ····················· | ···················· | ···················· | ···················· |
| J | 5892 | hCoV-19/England/QEUH-9B8E13/2020\|EPI_ISL_549663\|20 | ···················· | ···················· | ····················· | ···················· | ···················· | ···················· |
| J | 5893 | hCoV-19/England/QEUH-9B855A/2020\|EPI_ISL_549664\|20 | ···················· | ···················· | ····················· | ···················· | ···················· | ···················· |
| J | 5894 | hCoV-19/England/QEUH-9B9AC3/2020\|EPI_ISL_549665\|20 | ···················· | ···················· | ····················· | ···················· | ···················· | ···················· |
| J | 5895 | hCoV-19/England/QEUH-9B8E9B/2020\|EPI_ISL_549666\|20 | ···················· | ···················· | ····················· | ···················· | ···················· | ···················· |
| J | 5896 | hCoV-19/England/QEUH-9B84A8/2020\|EPI_ISL_549667\|20 | ···················· | ···················· | ····················· | ···················· | ···················· | ···················· |
| J | 5897 | hCoV-19/England/QEUH-9B9F1F/2020\|EPI_ISL_549668\|20 | ···················· | ···················· | ····················· | ···················· | ···················· | ···················· |
| J | 5898 | hCoV-19/England/QEUH-9B8D43/2020\|EPI_ISL_549669\|20 | ···················· | ·A·················· | ····················· | ···················· | ···················· | ···················· |
| J | 5899 | hCoV-19/England/MILK-99C862/2020\|EPI_ISL_550640\|20 | ···················· | ···················· | ····················· | ···················· | ···················· | ···················· |
| J | 5900 | hCoV-19/England/MILK-99E1DE/2020\|EPI_ISL_550642\|20 | ···················· | ···················· | ····················· | ···················· | ···················· | ···················· |
| J | 5901 | hCoV-19/England/MILK-99C792/2020\|EPI_ISL_550643\|20 | ···················· | ···················· | ····················· | ···················· | ···················· | ···················· |
| J | 5902 | hCoV-19/England/MILK-99C440/2020\|EPI_ISL_550644\|20 | ···················· | ···················· | ····················· | ···················· | ···················· | ···················· |
| J | 5903 | hCoV-19/England/MILK-99DC83/2020\|EPI_ISL_550645\|20 | ···················· | ···················· | ····················· | ···················· | ···················· | ···················· |
| J | 5904 | hCoV-19/Wales/MILK-99CA8A/2020\|EPI_ISL_550646\|2020 | ···················· | ···················· | ····················· | ···················· | ···················· | ···················· |
| J | 5905 | hCoV-19/England/MILK-99DE05/2020\|EPI_ISL_550647\|20 | ···················· | ···················· | ····················· | ···················· | ···················· | ···················· |
| J | 5906 | hCoV-19/England/MILK-99E5A5/2020\|EPI_ISL_550648\|20 | ···················· | ···················· | ····················· | ···················· | ···················· | ···················· |
| J | 5907 | hCoV-19/England/MILK-99D421/2020\|EPI_ISL_550649\|20 | ···················· | ···················· | ····················· | ···················· | ···················· | ···················· |
| J | 5908 | hCoV-19/England/MILK-99DD17/2020\|EPI_ISL_550650\|20 | ···················· | ···················· | ····················· | ···················· | ···················· | ···················· |
| J | 5909 | hCoV-19/England/MILK-991E98/2020\|EPI_ISL_551972\|20 | ···················· | ···················· | ····················· | ···················· | ···················· | ···················· |
| J | 5910 | hCoV-19/Wales/MILK-9936BC/2020\|EPI_ISL_551974\|2020 | ···················· | ···················· | ····················· | ···················· | ···················· | ···················· |
| J | 5911 | hCoV-19/England/MILK-993054/2020\|EPI_ISL_551975\|20 | ···················· | ···················· | ····················· | ···················· | ···················· | ···················· |
| J | 5912 | hCoV-19/England/MILK-99286C/2020\|EPI_ISL_551976\|20 | ···················· | ···················· | ····················· | ···················· | ···················· | ···················· |
| J | 5913 | hCoV-19/England/MILK-992E00/2020\|EPI_ISL_551978\|20 | ···················· | ···················· | ····················· | ···················· | ···················· | ···················· |

J 5914 hCoV-19/England/MILK-992750/2020|EPI_ISL_551979|20 ···················· ···················· ····················· ···················· ···················· ····················
J 5915 hCoV-19/England/CAMC-9916CD/2020|EPI_ISL_551980|20 ···················· ···················· ····················· ···················· ···················· ····················
J 5916 hCoV-19/England/MILK-992D9A/2020|EPI_ISL_551981|20 ···················· ···················· ····················· ···················· ···················· ····················
J 5917 hCoV-19/England/MILK-957BF1/2020|EPI_ISL_554127|20 ···················· ···················· ····················· ···················· ···················· ····················
J 5918 hCoV-19/England/MILK-956FE7/2020|EPI_ISL_554128|20 ···················· ···················· ····················· ···················· ···················· ····················
J 5919 hCoV-19/England/ALDP-951368/2020|EPI_ISL_554129|20 ···················· ···················· ····················· ···················· ···················· ····················
J 5920 hCoV-19/England/ALDP-950EC2/2020|EPI_ISL_554130|20 ···················· ···················· ····················· ···················· ···················· ····················
J 5921 hCoV-19/England/MILK-956FC9/2020|EPI_ISL_554132|20 ···················· ···················· ····················· ···················· ···················· ····················
J 5922 hCoV-19/England/MILK-9579BB/2020|EPI_ISL_554133|20 ···················· ···················· ····················· ···················· ···················· ····················
J 5923 hCoV-19/England/MILK-957BC4/2020|EPI_ISL_554138|20 ···················· ···················· ····················· ···················· ···················· ····················
J 5924 hCoV-19/England/MILK-95797F/2020|EPI_ISL_554139|20 ···················· ···················· ····················· ···················· ···················· ····················
J 5925 hCoV-19/Wales/MILK-956916/2020|EPI_ISL_554142|2020 ···················· ···················· ····················· ···················· ···················· ····················
J 5926 hCoV-19/England/MILK-957283/2020|EPI_ISL_554143|20 ···················· ···················· ····················· ···················· ···················· ····················
J 5927 hCoV-19/England/MILK-956A22/2020|EPI_ISL_554144|20 ···················· ···················· ····················· ···················· ···················· ····················
J 5928 hCoV-19/England/MILK-956AF5/2020|EPI_ISL_554146|20 ···················· ···················· ····················· ···················· ···················· ····················
J 5929 hCoV-19/England/MILK-9567FE/2020|EPI_ISL_554147|20 ···················· ···················· ····················· ···················· ···················· ····················
J 5930 hCoV-19/England/MILK-95702F/2020|EPI_ISL_554148|20 ···················· ···················· ····················· ···················· ···················· ····················
J 5931 hCoV-19/England/MILK-956891/2020|EPI_ISL_554150|20 ···················· ···················· ····················· ···················· ···················· ····················
J 5932 hCoV-19/England/MILK-957AD6/2020|EPI_ISL_554152|20 ···················· ···················· ····················· ···················· ···················· ····················
J 5933 hCoV-19/England/MILK-956B5C/2020|EPI_ISL_554153|20 ···················· ···················· ····················· ···················· ···················· ····················
J 5934 hCoV-19/England/MILK-957B88/2020|EPI_ISL_554154|20 ···················· ····A··············· ····················· ···················· ···················· ····················
J 5935 hCoV-19/England/ALDP-9512F2/2020|EPI_ISL_554155|20 ···················· ···················· ····················· ···················· ···················· ····················
J 5936 hCoV-19/England/ALDP-9512E3/2020|EPI_ISL_554156|20 ···················· ·A·················· ····················· ···················· ···················· ····················
J 5937 hCoV-19/Wales/ALDP-950F56/2020|EPI_ISL_554157|2020 ···················· ···················· ····················· ···················· ···················· ····················
J 5938 hCoV-19/England/ALDP-951016/2020|EPI_ISL_554158|20 ···················· ···················· ····················· ···················· ···················· ····················
J 5939 hCoV-19/England/MILK-95706B/2020|EPI_ISL_554159|20 ···················· ···················· ····················· ···················· ···················· ····················
J 5940 hCoV-19/England/MILK-957BA6/2020|EPI_ISL_554160|20 ···················· ·A·················· ····················· ···················· ···················· ····················
J 5941 hCoV-19/England/MILK-956B6B/2020|EPI_ISL_554163|20 ···················· ···················· ····················· ···················· ···················· ····················
J 5942 hCoV-19/Wales/ALDP-951359/2020|EPI_ISL_554164|2020 ···················· ···················· ····················· ···················· ···················· ····················
J 5943 hCoV-19/England/ALDP-950FA1/2020|EPI_ISL_554165|20 ···················· ···················· ····················· ···················· ···················· ····················
J 5944 hCoV-19/England/MILK-956C59/2020|EPI_ISL_554166|20 ···················· ···················· ····················· ···················· ···················· ····················
J 5945 hCoV-19/England/MILK-957B5B/2020|EPI_ISL_554168|20 ···················· ···················· ····················· ···················· ···················· ····················
J 5946 hCoV-19/England/MILK-957344/2020|EPI_ISL_554169|20 ···················· ···················· ····················· ···················· ···················· ····················
J 5947 hCoV-19/England/MILK-957B3D/2020|EPI_ISL_554200|20 ···················· ···················· ····················· ···················· ···················· ····················
J 5948 hCoV-19/England/MILK-956794/2020|EPI_ISL_554201|20 ···················· ···················· ····················· ···················· ···················· ····················
J 5949 hCoV-19/England/ALDP-951034/2020|EPI_ISL_554202|20 ···················· ···················· ····················· ···················· ···················· ····················
J 5950 hCoV-19/England/ALDP-95134A/2020|EPI_ISL_554203|20 ···················· ···················· ····················· ···················· ···················· ····················
J 5951 hCoV-19/England/ALDP-954839/2020|EPI_ISL_555458|20 ···················· ···················· ····················· ···················· ···················· ····················
J 5952 hCoV-19/Wales/ALDP-952C8A/2020|EPI_ISL_555460|2020 ···················· ···················· ····················· ···················· ···················· ····················
J 5953 hCoV-19/England/ALDP-952B50/2020|EPI_ISL_555461|20 ···················· ···················· ····················· ···················· ···················· ····················
J 5954 hCoV-19/England/ALDP-94DFE3/2020|EPI_ISL_555462|20 ···················· ···················· ····················· ···················· ···················· ····················
J 5955 hCoV-19/England/ALDP-94DFF2/2020|EPI_ISL_555463|20 ···················· ···················· ····················· ···················· ···················· ····················
J 5956 hCoV-19/England/ALDP-9547E1/2020|EPI_ISL_555465|20 ···················· ···················· ····················· ···················· ···················· ····················
J 5957 hCoV-19/England/ALDP-951878/2020|EPI_ISL_555469|20 ···················· ·A·················· ····················· ···················· ···················· ····················
J 5958 hCoV-19/England/ALDP-95181E/2020|EPI_ISL_555470|20 ···················· ···················· ····················· ···················· ···················· ····················
J 5959 hCoV-19/England/ALDP-94E5B3/2020|EPI_ISL_555473|20 ···················· ···················· ····················· ···················· ···················· ····················
J 5960 hCoV-19/England/ALDP-9547A5/2020|EPI_ISL_555476|20 ···················· ···················· ····················· ···················· ···················· ····················
J 5961 hCoV-19/England/ALDP-951C21/2020|EPI_ISL_555477|20 ···················· ···················· ····················· ···················· ···················· ····················
J 5962 hCoV-19/England/ALDP-94E8D8/2020|EPI_ISL_555478|20 ···················· ···················· ····················· ···················· ···················· ····················
J 5963 hCoV-19/England/ALDP-94E076/2020|EPI_ISL_555479|20 ···················· ···················· ····················· ···················· ···················· ····················
J 5964 hCoV-19/England/ALDP-94E54A/2020|EPI_ISL_555480|20 ···················· ···················· ····················· ···················· ···················· ····················
J 5965 hCoV-19/England/ALDP-951975/2020|EPI_ISL_555481|20 ···················· ···················· ····················· ···················· ···················· ····················
J 5966 hCoV-19/England/ALDP-955C1E/2020|EPI_ISL_555482|20 ···················· ···················· ····················· ···················· ···················· ····················
J 5967 hCoV-19/England/ALDP-9559EA/2020|EPI_ISL_555483|20 ···················· ···················· ····················· ···················· ···················· ····················
J 5968 hCoV-19/England/ALDP-94E98A/2020|EPI_ISL_555484|20 ···················· ···················· ····················· ···················· ···················· ····················
J 5969 hCoV-19/England/ALDP-95184B/2020|EPI_ISL_555485|20 ···················· ···················· ····················· ···················· ···················· ····················
J 5970 hCoV-19/England/ALDP-94E067/2020|EPI_ISL_555486|20 ···················· ···················· ····················· ···················· ···················· ····················
J 5971 hCoV-19/England/ALDP-94E6A1/2020|EPI_ISL_555487|20 ···················· ·A·················· ····················· ···················· ···················· ····················
J 5972 hCoV-19/England/ALDP-9517B7/2020|EPI_ISL_555488|20 ···················· ·A·················· ····················· ···················· ···················· ····················
J 5973 hCoV-19/England/ALDP-955BB7/2020|EPI_ISL_555489|20 ···················· ···················· ····················· ···················· ···················· ····················
J 5974 hCoV-19/England/ALDP-95192A/2020|EPI_ISL_555490|20 ···················· ···················· ····················· ···················· ···················· ····················
J 5975 hCoV-19/England/ALDP-94E8C9/2020|EPI_ISL_555491|20 ···················· ···················· ····················· ···················· ···················· ····················
J 5976 hCoV-19/England/ALDP-9559CC/2020|EPI_ISL_555492|20 ···················· ···················· ····················· ···················· ···················· ····················
J 5977 hCoV-19/England/ALDP-954866/2020|EPI_ISL_555494|20 ···················· ···················· ····················· ···················· ···················· ····················
J 5978 hCoV-19/England/ALDP-9517F3/2020|EPI_ISL_555495|20 ···················· ···················· ····················· ···················· ···················· ····················
J 5979 hCoV-19/England/ALDP-951984/2020|EPI_ISL_555496|20 ···················· ···················· ····················· ···················· ···················· ····················
J 5980 hCoV-19/England/ALDP-954909/2020|EPI_ISL_555497|20 ···················· ···················· ····················· ···················· ···················· ····················
J 5981 hCoV-19/England/ALDP-94E094/2020|EPI_ISL_555498|20 ···················· ···················· ····················· ···················· ···················· ····················
J 5982 hCoV-19/England/ALDP-94E47A/2020|EPI_ISL_555499|20 ···················· ···················· ····················· ···················· ···················· ····················
J 5983 hCoV-19/England/ALDP-955A14/2020|EPI_ISL_555501|20 ···················· ···················· ····················· ···················· ···················· ····················
J 5984 hCoV-19/England/ALDP-94E3B9/2020|EPI_ISL_555503|20 ···················· ···················· ····················· ···················· ···················· ····················
J 5985 hCoV-19/England/ALDP-951A90/2020|EPI_ISL_555504|20 ···················· ···················· ····················· ···················· ···················· ····················
J 5986 hCoV-19/England/ALDP-94DE9B/2020|EPI_ISL_555506|20 ···················· ···················· ····················· ···················· ···················· ····················
J 5987 hCoV-19/England/ALDP-944B9D/2020|EPI_ISL_555507|20 ···················· ···················· ····················· ···················· ···················· ····················
J 5988 hCoV-19/England/ALDP-95AF7A/2020|EPI_ISL_555508|20 ···················· ···················· ····················· ···················· ···················· ····················
J 5989 hCoV-19/England/ALDP-663032/2020|EPI_ISL_555509|20 ···················· ···················· ····················· ···················· ···················· ····················
J 5990 hCoV-19/England/ALDP-9448C3/2020|EPI_ISL_555512|20 ···················· ···················· ····················· ···················· ···················· ····················
J 5991 hCoV-19/England/ALDP-94DE6E/2020|EPI_ISL_555513|20 ···················· ···················· ····················· ···················· ···················· ····················
J 5992 hCoV-19/England/ALDP-95240A/2020|EPI_ISL_555514|20 ···················· ···················· ····················· ···················· ···················· ····················
J 5993 hCoV-19/England/ALDP-661975/2020|EPI_ISL_555515|20 ···················· ···················· ····················· ···················· ···················· ····················

| | | | | | | | | |
|---|---|---|---|---|---|---|---|---|
| J | 5994 | hCoV-19/England/ALDP-95ADBC/2020\|EPI_ISL_555516\|20 | ···················· | ···················· | ····················· | ···················· | ···················· | ···················· |
| J | 5995 | hCoV-19/England/ALDP-94DEB9/2020\|EPI_ISL_555517\|20 | ···················· | ···················· | ····················· | ···················· | ···················· | ···················· |
| J | 5996 | hCoV-19/England/ALDP-65D94F/2020\|EPI_ISL_555518\|20 | ···················· | ···················· | ····················· | ···················· | ···················· | ···················· |
| J | 5997 | hCoV-19/England/ALDP-95ACDD/2020\|EPI_ISL_555520\|20 | ···················· | ···················· | ····················· | ···················· | ···················· | ···················· |
| J | 5998 | hCoV-19/England/ALDP-94491B/2020\|EPI_ISL_555522\|20 | ···················· | ···················· | ····················· | ···················· | ···················· | ···················· |
| J | 5999 | hCoV-19/England/ALDP-95AB49/2020\|EPI_ISL_555523\|20 | ···················· | ···················· | ····················· | ···················· | ···················· | ···················· |
| J | 6000 | hCoV-19/England/ALDP-94DC37/2020\|EPI_ISL_555524\|20 | ···················· | ···················· | ····················· | ···················· | ···················· | ···················· |
| J | 6001 | hCoV-19/England/ALDP-9446D8/2020\|EPI_ISL_555525\|20 | ···················· | ···················· | ····················· | ···················· | ···················· | ···················· |
| J | 6002 | hCoV-19/England/ALDP-95B137/2020\|EPI_ISL_555526\|20 | ···················· | ···················· | ····················· | ···················· | ···················· | ···················· |
| J | 6003 | hCoV-19/England/ALDP-95B155/2020\|EPI_ISL_555527\|20 | ···················· | ···················· | ····················· | ···················· | ···················· | ···················· |
| J | 6004 | hCoV-19/England/ALDP-664A15/2020\|EPI_ISL_555528\|20 | ···················· | ···················· | ····················· | ···················· | ···················· | ···················· |
| J | 6005 | hCoV-19/England/ALDP-95AD8F/2020\|EPI_ISL_555529\|20 | ···················· | ···················· | ····················· | ···················· | ···················· | ···················· |
| J | 6006 | hCoV-19/England/ALDP-6675E4/2020\|EPI_ISL_555530\|20 | ···················· | ···················· | ····················· | ···················· | ···················· | ···················· |
| J | 6007 | hCoV-19/England/ALDP-95B094/2020\|EPI_ISL_555531\|20 | ···················· | ···················· | ····················· | ···················· | ···················· | ···················· |
| J | 6008 | hCoV-19/England/ALDP-944BD9/2020\|EPI_ISL_555532\|20 | ···················· | ···················· | ····················· | ···················· | ···················· | ···················· |
| J | 6009 | hCoV-19/England/ALDP-95B00D/2020\|EPI_ISL_555533\|20 | ···················· | ···················· | ····················· | ···················· | ···················· | ···················· |
| J | 6010 | hCoV-19/England/ALDP-94476C/2020\|EPI_ISL_555534\|20 | ···················· | ···················· | ····················· | ···················· | ···················· | ···················· |
| J | 6011 | hCoV-19/England/CAMC-945507/2020\|EPI_ISL_556789\|20 | ···················· | ···················· | ····················· | ···················· | ···················· | ···················· |
| J | 6012 | hCoV-19/England/CAMC-945473/2020\|EPI_ISL_556790\|20 | ···················· | ···················· | ····················· | ···················· | ···················· | ···················· |
| J | 6013 | hCoV-19/England/CAMC-94B792/2020\|EPI_ISL_556792\|20 | ···················· | ······A············· | ····················· | ···················· | ···················· | ···················· |
| J | 6014 | hCoV-19/England/CAMC-9451A9/2020\|EPI_ISL_556794\|20 | ···················· | ···················· | ····················· | ···················· | ···················· | ···················· |
| J | 6015 | hCoV-19/England/CAMC-946DC2/2020\|EPI_ISL_556795\|20 | ···················· | ···················· | ····················· | ···················· | ···················· | ···················· |
| J | 6016 | hCoV-19/England/CAMC-945640/2020\|EPI_ISL_556796\|20 | ···················· | ···················· | ····················· | ···················· | ···················· | ···················· |
| J | 6017 | hCoV-19/England/CAMC-945947/2020\|EPI_ISL_556801\|20 | ···················· | ···················· | ····················· | ···················· | ···················· | ···················· |
| J | 6018 | hCoV-19/England/CAMC-94AC94/2020\|EPI_ISL_556802\|20 | ···················· | ···················· | ····················· | ···················· | ···················· | ···················· |
| J | 6019 | hCoV-19/England/CAMC-94B176/2020\|EPI_ISL_556804\|20 | ···················· | ···················· | ····················· | ···················· | ···················· | ···················· |
| J | 6020 | hCoV-19/England/CAMC-948522/2020\|EPI_ISL_556805\|20 | ···················· | ···················· | ····················· | ···················· | ···················· | ···················· |
| J | 6021 | hCoV-19/England/CAMC-94BA02/2020\|EPI_ISL_556806\|20 | ···················· | ······A············· | ····················· | ···················· | ···················· | ···················· |
| J | 6022 | hCoV-19/England/CAMC-946CC5/2020\|EPI_ISL_556807\|20 | ···················· | ···················· | ····················· | ···················· | ···················· | ···················· |
| J | 6023 | hCoV-19/England/CAMC-94956D/2020\|EPI_ISL_556808\|20 | ···················· | ·A·················· | ····················· | ···················· | ···················· | ···················· |
| J | 6024 | hCoV-19/England/CAMC-9485C8/2020\|EPI_ISL_556809\|20 | ···················· | ···················· | ····················· | ···················· | ···················· | ···················· |
| J | 6025 | hCoV-19/England/CAMC-94881A/2020\|EPI_ISL_556810\|20 | ···················· | ···················· | ····················· | ···················· | ···················· | ···················· |
| J | 6026 | hCoV-19/England/CAMC-948513/2020\|EPI_ISL_556811\|20 | ···················· | ···················· | ····················· | ···················· | ···················· | ···················· |
| J | 6027 | hCoV-19/England/CAMC-9456B9/2020\|EPI_ISL_556812\|20 | ···················· | ···················· | ····················· | ···················· | ···················· | ···················· |
| J | 6028 | hCoV-19/England/CAMC-946C7A/2020\|EPI_ISL_556813\|20 | ···················· | ···················· | ····················· | ···················· | ···················· | ···················· |
| J | 6029 | hCoV-19/England/CAMC-9458C2/2020\|EPI_ISL_556814\|20 | ···················· | ···················· | ····················· | ···················· | ···················· | ···················· |
| J | 6030 | hCoV-19/England/CAMC-946D86/2020\|EPI_ISL_556815\|20 | ···················· | ···················· | ····················· | ···················· | ···················· | ···················· |
| J | 6031 | hCoV-19/England/CAMC-94915A/2020\|EPI_ISL_556816\|20 | ···················· | ···················· | ····················· | ···················· | ···················· | ···················· |
| J | 6032 | hCoV-19/England/CAMC-945868/2020\|EPI_ISL_556818\|20 | ···················· | ···················· | ····················· | ···················· | ···················· | ···················· |
| J | 6033 | hCoV-19/England/CAMC-94C078/2020\|EPI_ISL_556819\|20 | ···················· | ·A·················· | ····················· | ···················· | ···················· | ···················· |
| J | 6034 | hCoV-19/England/CAMC-946C89/2020\|EPI_ISL_556820\|20 | ···················· | ···················· | ····················· | ···················· | ···················· | ···················· |
| J | 6035 | hCoV-19/England/CAMC-94857D/2020\|EPI_ISL_556821\|20 | ···················· | ·A·················· | ····················· | ···················· | ···················· | ···················· |
| J | 6036 | hCoV-19/England/CAMC-948BF3/2020\|EPI_ISL_556823\|20 | ···················· | ···················· | ····················· | ···················· | ···················· | ···················· |
| J | 6037 | hCoV-19/England/CAMC-941635/2020\|EPI_ISL_556825\|20 | ···················· | ···················· | ····················· | ···················· | ···················· | ···················· |
| J | 6038 | hCoV-19/England/CAMC-941A1B/2020\|EPI_ISL_556827\|20 | ···················· | ···················· | ····················· | ···················· | ···················· | ···················· |
| J | 6039 | hCoV-19/England/CAMC-9414FF/2020\|EPI_ISL_556830\|20 | ···················· | ···················· | ····················· | ···················· | ···················· | ···················· |
| J | 6040 | hCoV-19/England/CAMC-941A39/2020\|EPI_ISL_556831\|20 | ···················· | ···················· | ····················· | ···················· | ···················· | ···················· |
| J | 6041 | hCoV-19/England/CAMC-941D4F/2020\|EPI_ISL_556832\|20 | ···················· | ···················· | ····················· | ···················· | ···················· | ···················· |
| J | 6042 | hCoV-19/England/CAMC-94169F/2020\|EPI_ISL_556834\|20 | ···················· | ···················· | ····················· | ···················· | ···················· | ···················· |
| J | 6043 | hCoV-19/England/CAMC-941A66/2020\|EPI_ISL_556835\|20 | ···················· | ···················· | ····················· | ···················· | ···················· | ···················· |
| J | 6044 | hCoV-19/England/CAMC-9417AB/2020\|EPI_ISL_556837\|20 | ···················· | ···················· | ····················· | ···················· | ···················· | ···················· |
| J | 6045 | hCoV-19/England/CAMC-9418C6/2020\|EPI_ISL_556838\|20 | ···················· | ···················· | ····················· | ···················· | ···················· | ···················· |
| J | 6046 | hCoV-19/England/CAMC-941BAF/2020\|EPI_ISL_556839\|20 | ···················· | ···················· | ····················· | ···················· | ···················· | ···················· |
| J | 6047 | hCoV-19/England/CAMC-9418F3/2020\|EPI_ISL_556840\|20 | ···················· | ···················· | ····················· | ···················· | ···················· | ···················· |
| J | 6048 | hCoV-19/England/CAMC-94137A/2020\|EPI_ISL_556841\|20 | ···················· | ···················· | ····················· | ···················· | ···················· | ···················· |
| J | 6049 | hCoV-19/England/CAMC-941592/2020\|EPI_ISL_556842\|20 | ···················· | ···················· | ····················· | ···················· | ···················· | ···················· |
| J | 6050 | hCoV-19/England/CAMC-941C8E/2020\|EPI_ISL_556843\|20 | ···················· | ···················· | ····················· | ···················· | ···················· | ···················· |
| J | 6051 | hCoV-19/England/CAMC-941B18/2020\|EPI_ISL_556844\|20 | ···················· | ···················· | ····················· | ···················· | ···················· | ···················· |
| J | 6052 | hCoV-19/England/CAMC-9413B6/2020\|EPI_ISL_556845\|20 | ···················· | ·A·················· | ····················· | ···················· | ···················· | ···················· |
| J | 6053 | hCoV-19/England/CAMC-941987/2020\|EPI_ISL_556846\|20 | ···················· | ···················· | ····················· | ···················· | ···················· | ···················· |
| J | 6054 | hCoV-19/England/CAMC-9415B0/2020\|EPI_ISL_556847\|20 | ···················· | ······A············· | ····················· | ···················· | ···················· | ···················· |
| J | 6055 | hCoV-19/England/CAMC-941C60/2020\|EPI_ISL_556848\|20 | ···················· | ···················· | ····················· | ···················· | ···················· | ···················· |
| J | 6056 | hCoV-19/England/CAMC-941671/2020\|EPI_ISL_556849\|20 | ···················· | ···················· | ····················· | ···················· | ···················· | ···················· |
| J | 6057 | hCoV-19/England/CAMC-941C24/2020\|EPI_ISL_556850\|20 | ···················· | ···················· | ····················· | ···················· | ···················· | ···················· |
| J | 6058 | hCoV-19/England/CAMC-941D12/2020\|EPI_ISL_556851\|20 | ···················· | ···················· | ····················· | ···················· | ···················· | ···················· |
| J | 6059 | hCoV-19/England/CAMC-941529/2020\|EPI_ISL_556853\|20 | ···················· | ···················· | ····················· | ···················· | ···················· | ···················· |
| J | 6060 | hCoV-19/England/CAMC-9412F5/2020\|EPI_ISL_556854\|20 | ···················· | ···················· | ····················· | ···················· | ···················· | ···················· |
| J | 6061 | hCoV-19/England/CAMC-941A75/2020\|EPI_ISL_556855\|20 | ···················· | ···················· | ····················· | ···················· | ···················· | ···················· |
| J | 6062 | hCoV-19/England/CAMC-941899/2020\|EPI_ISL_556856\|20 | ···················· | ···················· | ····················· | ···················· | ···················· | ···················· |
| J | 6063 | hCoV-19/England/CAMC-941BBE/2020\|EPI_ISL_556858\|20 | ···················· | ···················· | ····················· | ···················· | ···················· | ···················· |
| J | 6064 | hCoV-19/England/CAMC-941398/2020\|EPI_ISL_556859\|20 | ···················· | ···················· | ····················· | ···················· | ···················· | ···················· |
| J | 6065 | hCoV-19/England/CAMC-941820/2020\|EPI_ISL_556860\|20 | ···················· | ···················· | ····················· | ···················· | ···················· | ···················· |
| J | 6066 | hCoV-19/England/CAMC-941608/2020\|EPI_ISL_556861\|20 | ···················· | ···················· | ····················· | ···················· | ···················· | ···················· |
| J | 6067 | hCoV-19/England/CAMC-941495/2020\|EPI_ISL_556862\|20 | ···················· | ···················· | ····················· | ···················· | ···················· | ···················· |
| J | 6068 | hCoV-19/England/CAMC-94286B/2020\|EPI_ISL_556864\|20 | ···················· | ···················· | ····················· | ···················· | ···················· | ···················· |
| J | 6069 | hCoV-19/England/CAMC-942968/2020\|EPI_ISL_556865\|20 | ···················· | ···················· | ····················· | ···················· | ···················· | ···················· |
| J | 6070 | hCoV-19/England/MILK-5097A3/2020\|EPI_ISL_559061\|20 | ···················· | ···················· | ····················· | ···················· | ···················· | ···················· |
| J | 6071 | hCoV-19/England/MILK-516D56/2020\|EPI_ISL_559063\|20 | ···················· | ···················· | ····················· | ···················· | ···················· | ···················· |
| J | 6072 | hCoV-19/England/ALDP-520032/2020\|EPI_ISL_559064\|20 | ···················· | ···················· | ····················· | ···················· | ···················· | ···················· |
| J | 6073 | hCoV-19/England/ALDP-52B996/2020\|EPI_ISL_559066\|20 | ···················· | ·A·················· | ····················· | ···················· | ···················· | ···················· |

| | | | | | | | | |
|---|---|---|---|---|---|---|---|---|
| J | 6074 | hCoV-19/England/ALDP-524AB8/2020\|EPI_ISL_559067\|20 | ···················· | ···················· | ····················· | ···················· | ···················· | ···················· |
| J | 6075 | hCoV-19/England/ALDP-5246E1/2020\|EPI_ISL_559068\|20 | ···················· | ···················· | ····················· | ···················· | ···················· | ···················· |
| J | 6076 | hCoV-19/England/ALDP-52B90F/2020\|EPI_ISL_559069\|20 | ···················· | ···················· | ····················· | ···················· | ···················· | ···················· |
| J | 6077 | hCoV-19/England/ALDP-52A636/2020\|EPI_ISL_559070\|20 | ···················· | ···················· | ····················· | ···················· | ···················· | ···················· |
| J | 6078 | hCoV-19/England/MILK-510EB3/2020\|EPI_ISL_559071\|20 | ···················· | ···················· | ····················· | ···················· | ···················· | ···················· |
| J | 6079 | hCoV-19/England/ALDP-526B0E/2020\|EPI_ISL_559072\|20 | ···················· | ···················· | ····················· | ···················· | ···················· | ···················· |
| J | 6080 | hCoV-19/England/ALDP-5262DC/2020\|EPI_ISL_559073\|20 | ···················· | ···················· | ····················· | ···················· | ···················· | ···················· |
| J | 6081 | hCoV-19/England/ALDP-523749/2020\|EPI_ISL_559074\|20 | ···················· | ···················· | ····················· | ···················· | ···················· | ···················· |
| J | 6082 | hCoV-19/England/ALDP-51DE4D/2020\|EPI_ISL_559075\|20 | ···················· | ···················· | ····················· | ···················· | ···················· | ···················· |
| J | 6083 | hCoV-19/England/ALDP-519CED/2020\|EPI_ISL_559076\|20 | ···················· | ···················· | ····················· | ···················· | ···················· | ···················· |
| J | 6084 | hCoV-19/England/ALDP-52AAA3/2020\|EPI_ISL_559078\|20 | ···················· | ···················· | ····················· | ···················· | ···················· | ···················· |
| J | 6085 | hCoV-19/England/ALDP-51D6FA/2020\|EPI_ISL_559079\|20 | ···················· | ···················· | ····················· | ···················· | ···················· | ···················· |
| J | 6086 | hCoV-19/England/MILK-510B52/2020\|EPI_ISL_559080\|20 | ···················· | ···················· | ····················· | ···················· | ···················· | ···················· |
| J | 6087 | hCoV-19/England/MILK-5135E8/2020\|EPI_ISL_559081\|20 | ···················· | ···················· | ····················· | ···················· | ···················· | ···················· |
| J | 6088 | hCoV-19/England/MILK-517B00/2020\|EPI_ISL_559084\|20 | ···················· | ···················· | ····················· | ···················· | ···················· | ···················· |
| J | 6089 | hCoV-19/England/ALDP-522829/2020\|EPI_ISL_559085\|20 | ···················· | ···················· | ····················· | ···················· | ···················· | ···················· |
| J | 6090 | hCoV-19/England/ALDP-5239BC/2020\|EPI_ISL_559086\|20 | ···················· | ···················· | ····················· | ···················· | ···················· | ···················· |
| J | 6091 | hCoV-19/England/ALDP-521EEC/2020\|EPI_ISL_559087\|20 | ···················· | ···················· | ····················· | ···················· | ···················· | ···················· |
| J | 6092 | hCoV-19/England/MILK-5127F2/2020\|EPI_ISL_559088\|20 | ···················· | ···················· | ····················· | ···················· | ···················· | ···················· |
| J | 6093 | hCoV-19/England/ALDP-520C10/2020\|EPI_ISL_559090\|20 | ···················· | ···················· | ····················· | ···················· | ···················· | ···················· |
| J | 6094 | hCoV-19/England/ALDP-522B8A/2020\|EPI_ISL_559091\|20 | ···················· | ···················· | ····················· | ···················· | ···················· | ···················· |
| J | 6095 | hCoV-19/England/ALDP-51F546/2020\|EPI_ISL_559093\|20 | ···················· | ···················· | ····················· | ···················· | ···················· | ···················· |
| J | 6096 | hCoV-19/England/ALDP-5201F3/2020\|EPI_ISL_559094\|20 | ···················· | ···················· | ····················· | ···················· | ···················· | ···················· |
| J | 6097 | hCoV-19/England/ALDP-527280/2020\|EPI_ISL_559096\|20 | ···················· | ···················· | ····················· | ···················· | ···················· | ···················· |
| J | 6098 | hCoV-19/England/ALDP-4F587B/2020\|EPI_ISL_559098\|20 | ···················· | ···················· | ····················· | ···················· | ···················· | ···················· |
| J | 6099 | hCoV-19/England/ALDP-500469/2020\|EPI_ISL_559099\|20 | ···················· | ···················· | ····················· | ···················· | ···················· | ···················· |
| J | 6100 | hCoV-19/Scotland/QEUH-9B0DE1/2020\|EPI_ISL_560045\|2 | ···················· | ···················· | ····················· | ···················· | ···················· | ···················· |
| J | 6101 | hCoV-19/Scotland/QEUH-9B0E48/2020\|EPI_ISL_560046\|2 | ···················· | ···················· | ····················· | ···················· | ···················· | ···················· |
| J | 6102 | hCoV-19/Scotland/QEUH-9B0E57/2020\|EPI_ISL_560047\|2 | ···················· | ···················· | ····················· | ···················· | ···················· | ···················· |
| J | 6103 | hCoV-19/Scotland/QEUH-9B0E75/2020\|EPI_ISL_560048\|2 | ···················· | ···················· | ····················· | ···················· | ···················· | ···················· |
| J | 6104 | hCoV-19/Scotland/QEUH-9B0EA2/2020\|EPI_ISL_560049\|2 | ···················· | ···················· | ····················· | ···················· | ···················· | ···················· |
| J | 6105 | hCoV-19/Scotland/QEUH-9B0EC0/2020\|EPI_ISL_560050\|2 | ···················· | ···················· | ····················· | ···················· | ···················· | ···················· |
| J | 6106 | hCoV-19/Scotland/QEUH-9B0F18/2020\|EPI_ISL_560051\|2 | ···················· | ···················· | ····················· | ···················· | ···················· | ···················· |
| J | 6107 | hCoV-19/Scotland/QEUH-9B0F27/2020\|EPI_ISL_560052\|2 | ···················· | ···················· | ····················· | ···················· | ···················· | ···················· |
| J | 6108 | hCoV-19/Scotland/QEUH-9B0F72/2020\|EPI_ISL_560053\|2 | ···················· | ···················· | ····················· | ···················· | ···················· | ···················· |
| J | 6109 | hCoV-19/Scotland/QEUH-9B3A50/2020\|EPI_ISL_560056\|2 | ···················· | ···················· | ····················· | ···················· | ···················· | ···················· |
| J | 6110 | hCoV-19/Scotland/QEUH-9B3B5D/2020\|EPI_ISL_560057\|2 | ···················· | ···················· | ····················· | ···················· | ···················· | ···················· |
| J | 6111 | hCoV-19/Scotland/QEUH-9B3B6C/2020\|EPI_ISL_560058\|2 | ···················· | ···················· | ····················· | ···················· | ···················· | ···················· |
| J | 6112 | hCoV-19/Scotland/QEUH-9B3C69/2020\|EPI_ISL_560060\|2 | ···················· | ···················· | ····················· | ···················· | ···················· | ···················· |
| J | 6113 | hCoV-19/Scotland/QEUH-9B3C78/2020\|EPI_ISL_560061\|2 | ···················· | ···················· | ····················· | ···················· | ···················· | ···················· |
| J | 6114 | hCoV-19/Scotland/QEUH-9B3D75/2020\|EPI_ISL_560063\|2 | ···················· | ···················· | ····················· | ···················· | ···················· | ···················· |
| J | 6115 | hCoV-19/Scotland/QEUH-9B3F24/2020\|EPI_ISL_560065\|2 | ···················· | ···················· | ····················· | ···················· | ···················· | ···················· |
| J | 6116 | hCoV-19/Scotland/QEUH-9B3F42/2020\|EPI_ISL_560066\|2 | ···················· | ···················· | ····················· | ···················· | ···················· | ···················· |
| J | 6117 | hCoV-19/Scotland/QEUH-9B3F51/2020\|EPI_ISL_560067\|2 | ···················· | ···················· | ····················· | ···················· | ···················· | ···················· |
| J | 6118 | hCoV-19/Scotland/QEUH-9B3F60/2020\|EPI_ISL_560068\|2 | ···················· | ···················· | ····················· | ···················· | ···················· | ···················· |
| J | 6119 | hCoV-19/Scotland/QEUH-9B4020/2020\|EPI_ISL_560069\|2 | ···················· | ···················· | ····················· | ···················· | ···················· | ···················· |
| J | 6120 | hCoV-19/Scotland/QEUH-9B40B7/2020\|EPI_ISL_560070\|2 | ···················· | ······C············· | ····················· | ···················· | ···················· | ···················· |
| J | 6121 | hCoV-19/Scotland/QEUH-9B40D5/2020\|EPI_ISL_560071\|2 | ···················· | ······C············· | ····················· | ···················· | ···················· | ···················· |
| J | 6122 | hCoV-19/Scotland/QEUH-9B4187/2020\|EPI_ISL_560072\|2 | ···················· | ···················· | ····················· | ···················· | ···················· | ···················· |
| J | 6123 | hCoV-19/Scotland/QEUH-9B41C3/2020\|EPI_ISL_560073\|2 | ···················· | ·A·················· | ····················· | ···················· | ···················· | ···················· |
| J | 6124 | hCoV-19/Scotland/QEUH-9B44F7/2020\|EPI_ISL_560074\|2 | ···················· | ···················· | ····················· | ···················· | ···················· | ···················· |
| J | 6125 | hCoV-19/Scotland/QEUH-9B4503/2020\|EPI_ISL_560075\|2 | ···················· | ···················· | ····················· | ···················· | ···················· | ···················· |
| J | 6126 | hCoV-19/Scotland/QEUH-9B4512/2020\|EPI_ISL_560076\|2 | ···················· | ···················· | ····················· | ···················· | ···················· | ···················· |
| J | 6127 | hCoV-19/Scotland/QEUH-9B4521/2020\|EPI_ISL_560077\|2 | ···················· | ···················· | ····················· | ···················· | ···················· | ···················· |
| J | 6128 | hCoV-19/Scotland/QEUH-9B4530/2020\|EPI_ISL_560078\|2 | ···················· | ···················· | ····················· | ···················· | ···················· | ···················· |
| J | 6129 | hCoV-19/Scotland/QEUH-9B454F/2020\|EPI_ISL_560079\|2 | ···················· | ···················· | ····················· | ···················· | ···················· | ···················· |
| J | 6130 | hCoV-19/Wales/MILK-9A88FA/2020\|EPI_ISL_560080\|2020 | ···················· | ···················· | ····················· | ···················· | ···················· | ···················· |
| J | 6131 | hCoV-19/Wales/MILK-9A89BB/2020\|EPI_ISL_560081\|2020 | ···················· | ···················· | ····················· | ···················· | ···················· | ···················· |
| J | 6132 | hCoV-19/Wales/MILK-9A89CA/2020\|EPI_ISL_560082\|2020 | ···················· | ···················· | ····················· | ···················· | ···················· | ···················· |
| J | 6133 | hCoV-19/Wales/PHWC-16A66B/2020\|EPI_ISL_560083\|2020 | ···················· | ···················· | ····················· | ···················· | ···················· | ···················· |
| J | 6134 | hCoV-19/Wales/PHWC-16A67A/2020\|EPI_ISL_560084\|2020 | ···················· | ···················· | ····················· | ···················· | ···················· | ···················· |
| J | 6135 | hCoV-19/Wales/PHWC-16A698/2020\|EPI_ISL_560086\|2020 | ···················· | ···················· | ····················· | ···················· | ···················· | ···················· |
| J | 6136 | hCoV-19/Wales/PHWC-16A6A7/2020\|EPI_ISL_560087\|2020 | ···················· | ···················· | ····················· | ···················· | ···················· | ···················· |
| J | 6137 | hCoV-19/Wales/PHWC-16A6B6/2020\|EPI_ISL_560088\|2020 | ···················· | ···················· | ····················· | ···················· | ···················· | ···················· |
| J | 6138 | hCoV-19/Wales/PHWC-16A6C5/2020\|EPI_ISL_560089\|2020 | ···················· | ···················· | ····················· | ···················· | ···················· | ···················· |
| J | 6139 | hCoV-19/Wales/PHWC-16A6D4/2020\|EPI_ISL_560090\|2020 | ···················· | ···················· | ····················· | ···················· | ···················· | ···················· |
| J | 6140 | hCoV-19/Wales/PHWC-16A71D/2020\|EPI_ISL_560094\|2020 | ···················· | ···················· | ····················· | ···················· | ···················· | ···················· |
| J | 6141 | hCoV-19/Wales/PHWC-16A72C/2020\|EPI_ISL_560095\|2020 | ···················· | ···················· | ····················· | ···················· | ···················· | ···················· |
| J | 6142 | hCoV-19/Wales/PHWC-16A74A/2020\|EPI_ISL_560097\|2020 | ···················· | ···················· | ····················· | ···················· | ···················· | ···················· |
| J | 6143 | hCoV-19/Wales/PHWC-16A759/2020\|EPI_ISL_560098\|2020 | ···················· | ···················· | ····················· | ···················· | ···················· | ···················· |
| J | 6144 | hCoV-19/Wales/PHWC-16A768/2020\|EPI_ISL_560099\|2020 | ···················· | ···················· | ····················· | ···················· | ···················· | ···················· |
| J | 6145 | hCoV-19/England/NOTT-113379/2020\|EPI_ISL_549500\|20 | ···················· | ·A·················· | ····················· | ···················· | ···················· | ···················· |
| J | 6146 | hCoV-19/England/NOTT-1133A6/2020\|EPI_ISL_549501\|20 | ···················· | ·A·················· | ····················· | ···················· | ···················· | ···················· |
| J | 6147 | hCoV-19/England/NOTT-1133C4/2020\|EPI_ISL_549502\|20 | ···················· | ···················· | ····················· | ···················· | ···················· | ···················· |
| J | 6148 | hCoV-19/England/NOTT-11341C/2020\|EPI_ISL_549503\|20 | ···················· | ···················· | ····················· | ···················· | ···················· | ···················· |
| J | 6149 | hCoV-19/England/NOTT-11342B/2020\|EPI_ISL_549504\|20 | ···················· | ·A·················· | ····················· | ···················· | ···················· | ···················· |
| J | 6150 | hCoV-19/England/NOTT-113449/2020\|EPI_ISL_549505\|20 | ···················· | ···················· | ····················· | ···················· | ···················· | ···················· |
| J | 6151 | hCoV-19/England/NOTT-113485/2020\|EPI_ISL_549506\|20 | ···················· | ·A·················· | ····················· | ···················· | ···················· | ···················· |
| J | 6152 | hCoV-19/England/QEUH-9B99F3/2020\|EPI_ISL_549670\|20 | ···················· | ···················· | ····················· | ···················· | ···················· | ···················· |
| J | 6153 | hCoV-19/England/QEUH-9B9DBB/2020\|EPI_ISL_549671\|20 | ···················· | ···················· | ····················· | ···················· | ···················· | ···················· |

| | | | | | | | | |
|---|---|---|---|---|---|---|---|---|
| J | 6154 | hCoV-19/England/QEUH-9B997B/2020\|EPI_ISL_549672\|20 | ···················· | ·A·················· | ···················· | ···················· | ···················· | ··A················· |
| J | 6155 | hCoV-19/England/QEUH-9B9B39/2020\|EPI_ISL_549673\|20 | ···················· | ···················· | ···················· | ···················· | ···················· | ···················· |
| J | 6156 | hCoV-19/England/QEUH-9B9BC0/2020\|EPI_ISL_549674\|20 | ···················· | ···················· | ···················· | ···················· | ···················· | ···················· |
| J | 6157 | hCoV-19/England/QEUH-9B8781/2020\|EPI_ISL_549675\|20 | ···················· | ···················· | ···················· | ···················· | ···················· | ···················· |
| J | 6158 | hCoV-19/England/QEUH-9B8E31/2020\|EPI_ISL_549676\|20 | ···················· | ···················· | ···················· | ···················· | ···················· | ···················· |
| J | 6159 | hCoV-19/England/QEUH-9B9049/2020\|EPI_ISL_549678\|20 | ···················· | ·A·················· | ···················· | ···················· | ···················· | ··A················· |
| J | 6160 | hCoV-19/England/QEUH-9B8A97/2020\|EPI_ISL_549679\|20 | ···················· | ···················· | ···················· | ···················· | ···················· | ···················· |
| J | 6161 | hCoV-19/England/QEUH-9B8B85/2020\|EPI_ISL_549680\|20 | ···················· | ·A·················· | ···················· | ···················· | ···················· | ··A················· |
| J | 6162 | hCoV-19/England/QEUH-9B89A9/2020\|EPI_ISL_549681\|20 | ···················· | ···················· | ···················· | ···················· | ···················· | ···················· |
| J | 6163 | hCoV-19/England/QEUH-9B9B57/2020\|EPI_ISL_549682\|20 | ···················· | ···················· | ···················· | ···················· | ····T··············· | ···················· |
| J | 6164 | hCoV-19/England/QEUH-9B8AA6/2020\|EPI_ISL_549684\|20 | ···················· | ···················· | ···················· | ···················· | ···················· | ···················· |
| J | 6165 | hCoV-19/England/QEUH-9B9067/2020\|EPI_ISL_549686\|20 | ···················· | ···················· | ···················· | ···················· | ···················· | ···················· |
| J | 6166 | hCoV-19/England/QEUH-9B9CCD/2020\|EPI_ISL_549687\|20 | ···················· | ·A·················· | ···················· | ···················· | ···················· | ··A················· |
| J | 6167 | hCoV-19/England/QEUH-9B8860/2020\|EPI_ISL_549688\|20 | ···················· | ·A·················· | ···················· | ···················· | ···················· | ···················· |
| J | 6168 | hCoV-19/England/QEUH-9B87EB/2020\|EPI_ISL_549689\|20 | ···················· | ···················· | ···················· | ···················· | ···················· | ···················· |
| J | 6169 | hCoV-19/England/QEUH-9B91FB/2020\|EPI_ISL_549690\|20 | ···················· | ···················· | ···················· | ···················· | ···················· | ···················· |
| J | 6170 | hCoV-19/England/QEUH-9B99B7/2020\|EPI_ISL_549691\|20 | ···················· | ···················· | ···················· | ···················· | ···················· | ···················· |
| J | 6171 | hCoV-19/England/QEUH-9B9E12/2020\|EPI_ISL_549692\|20 | ···················· | ··········A········· | ···················· | ···················· | ···················· | ···················· |
| J | 6172 | hCoV-19/England/QEUH-9B8B49/2020\|EPI_ISL_549693\|20 | ···················· | ···················· | ···················· | ···················· | ···················· | ···················· |
| J | 6173 | hCoV-19/England/QEUH-9B8C91/2020\|EPI_ISL_549694\|20 | ···················· | ···················· | ···················· | ···················· | ···················· | ···················· |
| J | 6174 | hCoV-19/England/QEUH-9B9DF7/2020\|EPI_ISL_549696\|20 | ···················· | ···················· | ···················· | ···················· | ···················· | ···················· |
| J | 6175 | hCoV-19/England/QEUH-9B83E7/2020\|EPI_ISL_549697\|20 | ···················· | ···················· | ···················· | ···················· | ···················· | ···················· |
| J | 6176 | hCoV-19/England/QEUH-9B900D/2020\|EPI_ISL_549698\|20 | ···················· | ···················· | ···················· | ···················· | ···················· | ···················· |
| J | 6177 | hCoV-19/England/QEUH-9B894F/2020\|EPI_ISL_549699\|20 | ···················· | ·A·················· | ···················· | ···················· | ···················· | ··A················· |
| J | 6178 | hCoV-19/England/MILK-99DDAE/2020\|EPI_ISL_550651\|20 | ···················· | ···················· | ···················· | ···················· | ···················· | ···················· |
| J | 6179 | hCoV-19/England/MILK-99C8BD/2020\|EPI_ISL_550652\|20 | ···················· | ···················· | ···················· | ···················· | ···················· | ···················· |
| J | 6180 | hCoV-19/England/MILK-99E4F3/2020\|EPI_ISL_550653\|20 | ···················· | ···················· | ···················· | ···················· | ···················· | ···················· |
| J | 6181 | hCoV-19/England/MILK-99DC56/2020\|EPI_ISL_550654\|20 | ···················· | ·A·················· | ···················· | ···················· | ···················· | ···················· |
| J | 6182 | hCoV-19/England/MILK-99E578/2020\|EPI_ISL_550656\|20 | ···················· | ·A·················· | ···················· | ···················· | ···················· | ···················· |
| J | 6183 | hCoV-19/England/MILK-99E2CC/2020\|EPI_ISL_550658\|20 | ···················· | ···················· | ···················· | ···················· | ···················· | ···················· |
| J | 6184 | hCoV-19/England/MILK-99E350/2020\|EPI_ISL_550659\|20 | ···················· | ···················· | ···················· | ···················· | ···················· | ···················· |
| J | 6185 | hCoV-19/England/MILK-99DBFF/2020\|EPI_ISL_550661\|20 | ···················· | ···················· | ···················· | ···················· | ···················· | ···················· |
| J | 6186 | hCoV-19/England/MILK-99C950/2020\|EPI_ISL_550662\|20 | ···················· | ···················· | ···················· | ···················· | ···················· | ···················· |
| J | 6187 | hCoV-19/England/MILK-99C71A/2020\|EPI_ISL_550663\|20 | ···················· | ···················· | ···················· | ···················· | ···················· | ···················· |
| J | 6188 | hCoV-19/England/MILK-99DF5D/2020\|EPI_ISL_550664\|20 | ···················· | ···················· | ···················· | ···················· | ······T············· | ···················· |
| J | 6189 | hCoV-19/Wales/MILK-99DA7A/2020\|EPI_ISL_550665\|2020 | ···················· | ···················· | ···················· | ···················· | ···················· | ···················· |
| J | 6190 | hCoV-19/England/MILK-99E55A/2020\|EPI_ISL_550666\|20 | ···················· | ···················· | ···················· | ···················· | ···················· | ···················· |
| J | 6191 | hCoV-19/England/MILK-99E04A/2020\|EPI_ISL_550667\|20 | ···················· | ·A·················· | ···················· | ···················· | ···················· | ··A················· |
| J | 6192 | hCoV-19/England/MILK-99C923/2020\|EPI_ISL_550668\|20 | ···················· | ···················· | ···················· | ···················· | ······T············· | ···················· |
| J | 6193 | hCoV-19/England/MILK-99DDDB/2020\|EPI_ISL_550670\|20 | ···················· | ···················· | ···················· | ···················· | ···················· | ···················· |
| J | 6194 | hCoV-19/England/MILK-99C7A1/2020\|EPI_ISL_550671\|20 | ···················· | ···················· | ···················· | ···················· | ······T············· | ···················· |
| J | 6195 | hCoV-19/England/MILK-99D430/2020\|EPI_ISL_550672\|20 | ···················· | ···················· | ···················· | ···················· | ···················· | ···················· |
| J | 6196 | hCoV-19/Wales/MILK-99DED8/2020\|EPI_ISL_550673\|2020 | ···················· | ···················· | ···················· | ···················· | ···················· | ···················· |
| J | 6197 | hCoV-19/England/MILK-99DF7B/2020\|EPI_ISL_550674\|20 | ···················· | ···················· | ···················· | ···················· | ···················· | ···················· |
| J | 6198 | hCoV-19/Wales/MILK-99DE8D/2020\|EPI_ISL_550675\|2020 | ···················· | ···················· | ···················· | ···················· | ···················· | ···················· |
| J | 6199 | hCoV-19/England/MILK-99DDEA/2020\|EPI_ISL_550676\|20 | ···················· | ···················· | ···················· | ···················· | ···················· | ···················· |
| J | 6200 | hCoV-19/England/MILK-99E226/2020\|EPI_ISL_550678\|20 | ···················· | ···················· | ···················· | ···················· | ···················· | ···················· |
| J | 6201 | hCoV-19/England/MILK-99CA3F/2020\|EPI_ISL_550679\|20 | ···················· | ···················· | ···················· | ···················· | ···················· | ···················· |
| J | 6202 | hCoV-19/England/MILK-99C404/2020\|EPI_ISL_550680\|20 | ···················· | ···················· | ···················· | ···················· | ···················· | ···················· |
| J | 6203 | hCoV-19/England/MILK-99D7EC/2020\|EPI_ISL_550681\|20 | ···················· | ···················· | ···················· | ···················· | ···················· | ···················· |
| J | 6204 | hCoV-19/Wales/MILK-99DA6B/2020\|EPI_ISL_550682\|2020 | ···················· | ···················· | ···················· | ···················· | ···················· | ···················· |
| J | 6205 | hCoV-19/England/MILK-99D2EB/2020\|EPI_ISL_550683\|20 | ···················· | ···················· | ···················· | ···················· | ···················· | ···················· |
| J | 6206 | hCoV-19/England/MILK-99DC65/2020\|EPI_ISL_550684\|20 | ···················· | ···················· | ···················· | ···················· | ···················· | ···················· |
| J | 6207 | hCoV-19/England/MILK-99E323/2020\|EPI_ISL_550685\|20 | ···················· | ···················· | ···················· | ···················· | ···················· | ···················· |
| J | 6208 | hCoV-19/England/MILK-99E1ED/2020\|EPI_ISL_550686\|20 | ···················· | ···················· | ···················· | ···················· | ···················· | ···················· |
| J | 6209 | hCoV-19/England/MILK-99E235/2020\|EPI_ISL_550687\|20 | ···················· | ···················· | ···················· | ···················· | ···················· | ···················· |
| J | 6210 | hCoV-19/England/MILK-99DD08/2020\|EPI_ISL_550688\|20 | ···················· | ···················· | ···················· | ···················· | ···················· | ···················· |
| J | 6211 | hCoV-19/England/MILK-99C695/2020\|EPI_ISL_550689\|20 | ···················· | ···················· | ···················· | ···················· | ···················· | ···················· |
| J | 6212 | hCoV-19/England/MILK-99DEC9/2020\|EPI_ISL_550690\|20 | ···················· | ···················· | ···················· | ···················· | ···················· | ···················· |
| J | 6213 | hCoV-19/England/MILK-99C6D1/2020\|EPI_ISL_550691\|20 | ···················· | ···················· | ···················· | ···················· | ······T············· | ···················· |
| J | 6214 | hCoV-19/England/MILK-99D852/2020\|EPI_ISL_550692\|20 | ···················· | ···················· | ···················· | ···················· | ···················· | ···················· |
| J | 6215 | hCoV-19/England/MILK-99CAB7/2020\|EPI_ISL_550693\|20 | ···················· | ···················· | ···················· | ···················· | ···················· | ···················· |
| J | 6216 | hCoV-19/England/MILK-99C422/2020\|EPI_ISL_550694\|20 | ···················· | ···················· | ···················· | ···················· | ···················· | ···················· |
| J | 6217 | hCoV-19/England/MILK-99C5D4/2020\|EPI_ISL_550695\|20 | ···················· | ······A············· | ···················· | ···················· | ···················· | ···················· |
| J | 6218 | hCoV-19/England/MILK-99DF3F/2020\|EPI_ISL_550696\|20 | ···················· | ···················· | ···················· | ···················· | ···················· | ···················· |
| J | 6219 | hCoV-19/England/MILK-99C8CC/2020\|EPI_ISL_550697\|20 | ···················· | ···················· | ···················· | ···················· | ···················· | ···················· |
| J | 6220 | hCoV-19/England/MILK-99E02C/2020\|EPI_ISL_550698\|20 | ···················· | ·A·················· | ···················· | ···················· | ···················· | ··A················· |
| J | 6221 | hCoV-19/England/MILK-99C4C8/2020\|EPI_ISL_550699\|20 | ···················· | ···················· | ···················· | ···················· | ···················· | ···················· |
| J | 6222 | hCoV-19/England/MILK-693A4A/2020\|EPI_ISL_551800\|20 | ···················· | ···················· | ···················· | ···················· | ···················· | ···················· |
| J | 6223 | hCoV-19/England/ALDP-689A74/2020\|EPI_ISL_551801\|20 | ···················· | ···················· | ···················· | ···················· | ···················· | ···················· |
| J | 6224 | hCoV-19/England/MILK-6A383E/2020\|EPI_ISL_551802\|20 | ···················· | ·A·················· | ···················· | ···················· | ···················· | ···················· |
| J | 6225 | hCoV-19/England/ALDP-68003F/2020\|EPI_ISL_551803\|20 | ···················· | ···················· | ···················· | ···················· | ···················· | ···················· |
| J | 6226 | hCoV-19/England/ALDP-67CB71/2020\|EPI_ISL_551804\|20 | ···················· | ···················· | ···················· | ···················· | ···················· | ···················· |
| J | 6227 | hCoV-19/England/ALDP-67F349/2020\|EPI_ISL_551806\|20 | ···················· | ·A·················· | ···················· | ···················· | ···················· | ···················· |
| J | 6228 | hCoV-19/England/ALDP-678A96/2020\|EPI_ISL_551807\|20 | ···················· | ···················· | ···················· | ···················· | ···················· | ···················· |
| J | 6229 | hCoV-19/England/ALDP-689959/2020\|EPI_ISL_551808\|20 | ···················· | ···················· | ···················· | ···················· | ···················· | ···················· |
| J | 6230 | hCoV-19/England/MILK-6A65F8/2020\|EPI_ISL_551809\|20 | ···················· | ···················· | ···················· | ···················· | ···················· | ···················· |
| J | 6231 | hCoV-19/England/ALDP-67CC41/2020\|EPI_ISL_551810\|20 | ···················· | ···················· | ···················· | ···················· | ···················· | ···················· |
| J | 6232 | hCoV-19/England/ALDP-681B3D/2020\|EPI_ISL_551811\|20 | ···················· | ···················· | ···················· | ···················· | ···················· | ···················· |
| J | 6233 | hCoV-19/England/MILK-6A7120/2020\|EPI_ISL_551814\|20 | ···················· | ···················· | ···················· | ···················· | ···················· | ···················· |

```
J    6234 hCoV-19/England/ALDP-689A65/2020|EPI_ISL_551815|20    ····················  ····················  ·····················  ····················  ····················  ····················
J    6235 hCoV-19/England/MILK-6ABF04/2020|EPI_ISL_551817|20    ····················  ····················  ·····················  ····················  ····················  ····················
J    6236 hCoV-19/England/ALDP-693682/2020|EPI_ISL_551818|20    ····················  ····················  ·····················  ····················  ····················  ····················
J    6237 hCoV-19/England/ALDP-6881BC/2020|EPI_ISL_551819|20    ····················  ····················  ·····················  ····················  ····················  ····················
J    6238 hCoV-19/England/MILK-99276F/2020|EPI_ISL_551973|20    ····················  ····················  ·····················  ····················  ····················  ····················
J    6239 hCoV-19/England/MILK-9929E1/2020|EPI_ISL_551982|20    ····················  ·A·······A··········  ·····················  ····················  ····················  ····················
J    6240 hCoV-19/England/CAMC-99137B/2020|EPI_ISL_551983|20    ····················  ····················  ·····················  ····················  ····················  ····················
J    6241 hCoV-19/England/MILK-9928F3/2020|EPI_ISL_551984|20    ····················  ····················  ·····················  ····················  ······T·············  ····················
J    6242 hCoV-19/England/MILK-992AC0/2020|EPI_ISL_551985|20    ····················  ·A··················  ·····················  ····················  ····················  ···A················
J    6243 hCoV-19/England/MILK-993397/2020|EPI_ISL_551986|20    ····················  ····················  ·····················  ····················  ····················  ····················
J    6244 hCoV-19/England/MILK-992D03/2020|EPI_ISL_551987|20    ····················  ····················  ·····················  ····················  ····················  ····················
J    6245 hCoV-19/England/MILK-992C24/2020|EPI_ISL_551989|20    ····················  ····················  ·····················  ····················  ····················  ····················
J    6246 hCoV-19/England/CAMC-9916BE/2020|EPI_ISL_551990|20    ····················  ····················  ·····················  ····················  ····················  ····················
J    6247 hCoV-19/England/MILK-99291E/2020|EPI_ISL_551991|20    ····················  ·A··················  ·····················  ····················  ····················  ···A················
J    6248 hCoV-19/England/MILK-99328B/2020|EPI_ISL_551992|20    ····················  ····················  ·····················  ····················  ····················  ····················
J    6249 hCoV-19/England/MILK-992F49/2020|EPI_ISL_551995|20    ····················  ····················  ·····················  ····················  ····················  ····················
J    6250 hCoV-19/England/MILK-992B72/2020|EPI_ISL_551996|20    ····················  ····················  ·····················  ····················  ····················  ····················
J    6251 hCoV-19/England/MILK-99376E/2020|EPI_ISL_551998|20    ····················  ····················  ·····················  ····················  ····················  ····················
J    6252 hCoV-19/England/MILK-99283F/2020|EPI_ISL_551999|20    ····················  ·A··················  ·····················  ····················  ····················  ···A················
J    6253 hCoV-19/England/ALDP-9564F7/2020|EPI_ISL_554050|20    ····················  ····················  ·····················  ····················  ····················  ····················
J    6254 hCoV-19/England/ALDP-952A71/2020|EPI_ISL_554051|20    ····················  ····················  ·····················  ····················  ····················  ····················
J    6255 hCoV-19/England/ALDP-9561F0/2020|EPI_ISL_554052|20    ····················  ····················  ·····················  ····················  ····················  ····················
J    6256 hCoV-19/England/ALDP-955FD9/2020|EPI_ISL_554053|20    ····················  ····················  ·····················  ····················  ····················  ····················
J    6257 hCoV-19/England/MILK-9578BE/2020|EPI_ISL_554054|20    ····················  ····················  ·····················  ····················  ····················  ····················
J    6258 hCoV-19/England/ALDP-9526C8/2020|EPI_ISL_554056|20    ····················  ····················  ·····················  ····················  ····················  ····················
J    6259 hCoV-19/England/MILK-9577EE/2020|EPI_ISL_554057|20    ····················  ·A··················  ·····················  ····················  ····················  ····················
J    6260 hCoV-19/England/MILK-957924/2020|EPI_ISL_554058|20    ····················  ·A··················  ·····················  ····················  ····················  ····················
J    6261 hCoV-19/England/ALDP-955E18/2020|EPI_ISL_554059|20    ····················  ····················  ·····················  ····················  ····················  ····················
J    6262 hCoV-19/England/ALDP-952516/2020|EPI_ISL_554060|20    ····················  ····················  ·····················  ····················  ····················  ····················
J    6263 hCoV-19/England/ALDP-95614B/2020|EPI_ISL_554061|20    ····················  ····················  ·····················  ····················  ····················  ····················
J    6264 hCoV-19/England/ALDP-952A17/2020|EPI_ISL_554062|20    ····················  ····················  ·····················  ····················  ····················  ····················
J    6265 hCoV-19/England/ALDP-9525E9/2020|EPI_ISL_554064|20    ····················  ····················  ·····················  ····················  ····················  ····················
J    6266 hCoV-19/England/MILK-9594D6/2020|EPI_ISL_554065|20    ····················  ····················  ·····················  ····················  ····················  ····················
J    6267 hCoV-19/England/ALDP-955DEE/2020|EPI_ISL_554066|20    ····················  ·A··················  ·····················  ····················  ····················  ····················
J    6268 hCoV-19/England/ALDP-956248/2020|EPI_ISL_554068|20    ····················  ·A··················  ·····················  ····················  ····················  ····················
J    6269 hCoV-19/England/ALDP-956354/2020|EPI_ISL_554069|20    ····················  ····················  ·····················  ····················  ····················  ····················
J    6270 hCoV-19/England/ALDP-95284A/2020|EPI_ISL_554070|20    ····················  ····················  ·····················  ····················  ····················  ····················
J    6271 hCoV-19/England/ALDP-955D2A/2020|EPI_ISL_554072|20    ····················  ····················  ·····················  ····················  ····················  ····················
J    6272 hCoV-19/England/ALDP-952877/2020|EPI_ISL_554073|20    ····················  ····················  ·····················  ····················  ····················  ····················
J    6273 hCoV-19/England/ALDP-955D1B/2020|EPI_ISL_554074|20    ····················  ····················  ·····················  ····················  ····················  ····················
J    6274 hCoV-19/England/ALDP-952A35/2020|EPI_ISL_554075|20    ····················  ····················  ·····················  ····················  ····················  ····················
J    6275 hCoV-19/England/ALDP-952A53/2020|EPI_ISL_554076|20    ····················  ····················  ·····················  ····················  ····················  ····················
J    6276 hCoV-19/England/ALDP-955E27/2020|EPI_ISL_554077|20    ····················  ·A··················  ·····················  ····················  ····················  ····················
J    6277 hCoV-19/England/ALDP-956424/2020|EPI_ISL_554078|20    ····················  ·A··················  ·····················  ····················  ····················  ····················
J    6278 hCoV-19/England/MILK-957687/2020|EPI_ISL_554080|20    ····················  ····················  ·····················  ····················  ····················  ····················
J    6279 hCoV-19/England/ALDP-952938/2020|EPI_ISL_554081|20    ····················  ····················  ·····················  ····················  ····················  ····················
J    6280 hCoV-19/England/ALDP-95611E/2020|EPI_ISL_554082|20    ····················  ····················  ·····················  ····················  ····················  ····················
J    6281 hCoV-19/England/ALDP-9564AC/2020|EPI_ISL_554083|20    ····················  ····················  ·····················  ····················  ····················  ····················
J    6282 hCoV-19/England/ALDP-9529DE/2020|EPI_ISL_554084|20    ····················  ····················  ·····················  ····················  ····················  ····················
J    6283 hCoV-19/England/ALDP-952A44/2020|EPI_ISL_554085|20    ····················  ····················  ·····················  ····················  ····················  ····················
J    6284 hCoV-19/England/ALDP-95659A/2020|EPI_ISL_554086|20    ····················  ····················  ·····················  ····················  ····················  ····················
J    6285 hCoV-19/England/ALDP-955D75/2020|EPI_ISL_554087|20    ····················  ····················  ·····················  ····················  ····················  ····················
J    6286 hCoV-19/England/ALDP-9565B8/2020|EPI_ISL_554090|20    ····················  ····················  ·····················  ····················  ····················  ····················
J    6287 hCoV-19/England/ALDP-955E81/2020|EPI_ISL_554091|20    ····················  ····················  ·····················  ····················  ····················  ····················
J    6288 hCoV-19/England/ALDP-952A62/2020|EPI_ISL_554092|20    ····················  ····················  ·····················  ····················  ····················  ····················
J    6289 hCoV-19/England/ALDP-9527D4/2020|EPI_ISL_554093|20    ····················  ····················  ·····················  ····················  ····················  ····················
J    6290 hCoV-19/England/ALDP-956266/2020|EPI_ISL_554094|20    ····················  ····················  ·····················  ····················  ····················  ····················
J    6291 hCoV-19/England/ALDP-9562A2/2020|EPI_ISL_554095|20    ····················  ····················  ·····················  ····················  ····················  ····················
J    6292 hCoV-19/England/ALDP-95655E/2020|EPI_ISL_554096|20    ····················  ····················  ·····················  ····················  ····················  ····················
J    6293 hCoV-19/England/MILK-95761E/2020|EPI_ISL_554097|20    ····················  ····················  ·····················  ····················  ····················  ····················
J    6294 hCoV-19/England/ALDP-9560B7/2020|EPI_ISL_554098|20    ····················  ····················  ·····················  ····················  ····················  ····················
J    6295 hCoV-19/England/MILK-9573DB/2020|EPI_ISL_554099|20    ····················  ····················  ·····················  ····················  ····················  ····················
J    6296 hCoV-19/England/ALDP-94E119/2020|EPI_ISL_555380|20    ····················  ····················  ·····················  ····················  ····················  ····················
J    6297 hCoV-19/England/ALDP-94E50E/2020|EPI_ISL_555381|20    ····················  ····················  ·····················  ····················  ····················  ····················
J    6298 hCoV-19/England/ALDP-94E410/2020|EPI_ISL_555382|20    ····················  ····················  ·····················  ····················  ····················  ····················
J    6299 hCoV-19/England/ALDP-94E6DE/2020|EPI_ISL_555383|20    ····················  ····················  ·····················  ····················  ····················  ····················
J    6300 hCoV-19/England/ALDP-94E89C/2020|EPI_ISL_555384|20    ····················  ····················  ·····················  ····················  ····················  ····················
J    6301 hCoV-19/England/ALDP-955A32/2020|EPI_ISL_555385|20    ····················  ····················  ·····················  ····················  ····················  ····················
J    6302 hCoV-19/England/ALDP-95191B/2020|EPI_ISL_555386|20    ····················  ·A··················  ·····················  ····················  ····················  ····················
J    6303 hCoV-19/England/ALDP-94E683/2020|EPI_ISL_555388|20    ····················  ····················  ·····················  ····················  ····················  ····················
J    6304 hCoV-19/England/ALDP-952C7B/2020|EPI_ISL_555389|20    ····················  ····················  ·····················  ····················  ····················  ····················
J    6305 hCoV-19/England/ALDP-955A05/2020|EPI_ISL_555390|20    ····················  ····················  ·····················  ····················  ····················  ····················
J    6306 hCoV-19/Wales/ALDP-94E568/2020|EPI_ISL_555392|2020    ····················  ····················  ·····················  ····················  ····················  ····················
J    6307 hCoV-19/England/ALDP-94DF6B/2020|EPI_ISL_555393|20    ····················  ····················  ·····················  ····················  ····················  ····················
J    6308 hCoV-19/England/ALDP-951ADC/2020|EPI_ISL_555394|20    ····················  ····················  ·····················  ····················  ····················  ····················
J    6309 hCoV-19/England/ALDP-94E049/2020|EPI_ISL_555395|20    ····················  ····················  ·····················  ····················  ····················  ····················
J    6310 hCoV-19/England/ALDP-94E586/2020|EPI_ISL_555397|20    ····················  ····················  ·····················  ····················  ····················  ····················
J    6311 hCoV-19/England/MILK-50A7FD/2020|EPI_ISL_559000|20    ····················  ····················  ·····················  ····················  ····················  ····················
J    6312 hCoV-19/England/ALDP-5284B6/2020|EPI_ISL_559001|20    ····················  ····················  ·····················  ····················  ····················  ····················
J    6313 hCoV-19/England/ALDP-52A627/2020|EPI_ISL_559002|20    ····················  ····················  ·····················  ····················  ····················  ····················
```

<table>
<tr><td>J</td><td>6314</td><td>hCoV-19/England/ALDP-522917/2020|EPI_ISL_559003|20</td><td>····················</td><td>····················</td><td>····················</td><td>····················</td><td>····················</td><td>····················</td><td></td></tr>
<tr><td>J</td><td>6315</td><td>hCoV-19/England/ALDP-519746/2020|EPI_ISL_559004|20</td><td>····················</td><td>····················</td><td>····················</td><td>····················</td><td>····················</td><td>····················</td><td></td></tr>
<tr><td>J</td><td>6316</td><td>hCoV-19/Wales/MILK-50D314/2020|EPI_ISL_559005|2020</td><td>····················</td><td>····················</td><td>····················</td><td>····················</td><td>····················</td><td>····················</td><td></td></tr>
<tr><td>J</td><td>6317</td><td>hCoV-19/England/ALDP-5276A2/2020|EPI_ISL_559006|20</td><td>····················</td><td>····················</td><td>····················</td><td>····················</td><td>····················</td><td>····················</td><td></td></tr>
<tr><td>J</td><td>6318</td><td>hCoV-19/England/ALDP-52B29B/2020|EPI_ISL_559007|20</td><td>····················</td><td>····················</td><td>····················</td><td>····················</td><td>····················</td><td>····················</td><td></td></tr>
<tr><td>J</td><td>6319</td><td>hCoV-19/England/ALDP-5297AD/2020|EPI_ISL_559008|20</td><td>····················</td><td>····················</td><td>····················</td><td>····················</td><td>····················</td><td>····················</td><td></td></tr>
<tr><td>J</td><td>6320</td><td>hCoV-19/Wales/MILK-511B15/2020|EPI_ISL_559009|2020</td><td>····················</td><td>····················</td><td>····················</td><td>····················</td><td>····················</td><td>····················</td><td></td></tr>
<tr><td>J</td><td>6321</td><td>hCoV-19/England/ALDP-521CE2/2020|EPI_ISL_559010|20</td><td>····················</td><td>····················</td><td>····················</td><td>····················</td><td>····················</td><td>····················</td><td></td></tr>
<tr><td>J</td><td>6322</td><td>hCoV-19/England/MILK-50A1D1/2020|EPI_ISL_559011|20</td><td>····················</td><td>····················</td><td>····················</td><td>····················</td><td>····················</td><td>····················</td><td></td></tr>
<tr><td>J</td><td>6323</td><td>hCoV-19/England/ALDP-528FA6/2020|EPI_ISL_559012|20</td><td>····················</td><td>····················</td><td>····················</td><td>····················</td><td>····················</td><td>····················</td><td></td></tr>
<tr><td>J</td><td>6324</td><td>hCoV-19/England/ALDP-519D17/2020|EPI_ISL_559014|20</td><td>····················</td><td>····················</td><td>····················</td><td>····················</td><td>····················</td><td>··················C·</td><td>1</td></tr>
<tr><td>J</td><td>6325</td><td>hCoV-19/England/ALDP-51FF75/2020|EPI_ISL_559015|20</td><td>····················</td><td>····················</td><td>····················</td><td>····················</td><td>····················</td><td>····················</td><td></td></tr>
<tr><td>J</td><td>6326</td><td>hCoV-19/England/ALDP-524E43/2020|EPI_ISL_559016|20</td><td>····················</td><td>····················</td><td>····················</td><td>····················</td><td>····················</td><td>····················</td><td></td></tr>
<tr><td>J</td><td>6327</td><td>hCoV-19/England/ALDP-5249BB/2020|EPI_ISL_559020|20</td><td>····················</td><td>····················</td><td>····················</td><td>····················</td><td>····················</td><td>····················</td><td></td></tr>
<tr><td>J</td><td>6328</td><td>hCoV-19/England/ALDP-526AA7/2020|EPI_ISL_559021|20</td><td>····················</td><td>····················</td><td>····················</td><td>····················</td><td>····················</td><td>····················</td><td></td></tr>
<tr><td>J</td><td>6329</td><td>hCoV-19/England/ALDP-522A23/2020|EPI_ISL_559023|20</td><td>····················</td><td>·A··················</td><td>····················</td><td>····················</td><td>····················</td><td>····················</td><td></td></tr>
<tr><td>J</td><td>6330</td><td>hCoV-19/England/ALDP-521D49/2020|EPI_ISL_559025|20</td><td>····················</td><td>····················</td><td>····················</td><td>····················</td><td>····················</td><td>····················</td><td></td></tr>
<tr><td>J</td><td>6331</td><td>hCoV-19/England/ALDP-52B125/2020|EPI_ISL_559027|20</td><td>····················</td><td>····················</td><td>····················</td><td>····················</td><td>····················</td><td>····················</td><td></td></tr>
<tr><td>J</td><td>6332</td><td>hCoV-19/England/MILK-5104B1/2020|EPI_ISL_559028|20</td><td>····················</td><td>····················</td><td>····················</td><td>····················</td><td>····················</td><td>····················</td><td></td></tr>
<tr><td>J</td><td>6333</td><td>hCoV-19/England/ALDP-51D44B/2020|EPI_ISL_559029|20</td><td>····················</td><td>····················</td><td>····················</td><td>····················</td><td>····················</td><td>····················</td><td></td></tr>
<tr><td>J</td><td>6334</td><td>hCoV-19/England/ALDP-51EF85/2020|EPI_ISL_559030|20</td><td>····················</td><td>····················</td><td>····················</td><td>····················</td><td>····················</td><td>····················</td><td></td></tr>
<tr><td>J</td><td>6335</td><td>hCoV-19/England/ALDP-52114D/2020|EPI_ISL_559031|20</td><td>····················</td><td>····················</td><td>····················</td><td>····················</td><td>····················</td><td>··················C·</td><td>1</td></tr>
<tr><td>J</td><td>6336</td><td>hCoV-19/England/ALDP-51F142/2020|EPI_ISL_559032|20</td><td>····················</td><td>····················</td><td>····················</td><td>····················</td><td>····················</td><td>····················</td><td></td></tr>
<tr><td>J</td><td>6337</td><td>hCoV-19/England/ALDP-52C230/2020|EPI_ISL_559035|20</td><td>····················</td><td>····················</td><td>····················</td><td>····················</td><td>····················</td><td>····················</td><td></td></tr>
<tr><td>J</td><td>6338</td><td>hCoV-19/England/ALDP-521893/2020|EPI_ISL_559036|20</td><td>····················</td><td>····················</td><td>····················</td><td>····················</td><td>····················</td><td>····················</td><td></td></tr>
<tr><td>J</td><td>6339</td><td>hCoV-19/England/ALDP-51FBAE/2020|EPI_ISL_559037|20</td><td>····················</td><td>····················</td><td>····················</td><td>····················</td><td>····················</td><td>····················</td><td></td></tr>
<tr><td>J</td><td>6340</td><td>hCoV-19/England/ALDP-524353/2020|EPI_ISL_559038|20</td><td>····················</td><td>····················</td><td>····················</td><td>····················</td><td>····················</td><td>····················</td><td></td></tr>
<tr><td>J</td><td>6341</td><td>hCoV-19/England/ALDP-523758/2020|EPI_ISL_559040|20</td><td>····················</td><td>····················</td><td>····················</td><td>····················</td><td>····················</td><td>····················</td><td></td></tr>
<tr><td>J</td><td>6342</td><td>hCoV-19/England/MILK-512376/2020|EPI_ISL_559043|20</td><td>····················</td><td>····················</td><td>····················</td><td>····················</td><td>····················</td><td>····················</td><td></td></tr>
<tr><td>J</td><td>6343</td><td>hCoV-19/England/ALDP-525A11/2020|EPI_ISL_559044|20</td><td>····················</td><td>····················</td><td>····················</td><td>····················</td><td>····················</td><td>····················</td><td></td></tr>
<tr><td>J</td><td>6344</td><td>hCoV-19/England/ALDP-52AA85/2020|EPI_ISL_559045|20</td><td>····················</td><td>····················</td><td>····················</td><td>····················</td><td>····················</td><td>····················</td><td></td></tr>
<tr><td>J</td><td>6345</td><td>hCoV-19/England/MILK-50DD70/2020|EPI_ISL_559047|20</td><td>····················</td><td>····················</td><td>····················</td><td>····················</td><td>····················</td><td>····················</td><td></td></tr>
<tr><td>J</td><td>6346</td><td>hCoV-19/Wales/MILK-5142FF/2020|EPI_ISL_559049|2020</td><td>····················</td><td>····················</td><td>····················</td><td>····················</td><td>····················</td><td>····················</td><td></td></tr>
<tr><td>J</td><td>6347</td><td>hCoV-19/England/ALDP-51F42B/2020|EPI_ISL_559050|20</td><td>····················</td><td>····················</td><td>····················</td><td>····················</td><td>····················</td><td>····················</td><td></td></tr>
<tr><td>J</td><td>6348</td><td>hCoV-19/England/ALDP-526209/2020|EPI_ISL_559051|20</td><td>····················</td><td>····················</td><td>····················</td><td>····················</td><td>····················</td><td>····················</td><td></td></tr>
<tr><td>J</td><td>6349</td><td>hCoV-19/England/ALDP-526E8D/2020|EPI_ISL_559052|20</td><td>····················</td><td>····················</td><td>····················</td><td>····················</td><td>····················</td><td>····················</td><td></td></tr>
<tr><td>J</td><td>6350</td><td>hCoV-19/England/ALDP-52B8D5/2020|EPI_ISL_559053|20</td><td>····················</td><td>····················</td><td>····················</td><td>····················</td><td>····················</td><td>····················</td><td></td></tr>
<tr><td>J</td><td>6351</td><td>hCoV-19/England/ALDP-51D4E1/2020|EPI_ISL_559054|20</td><td>····················</td><td>····················</td><td>····················</td><td>····················</td><td>····················</td><td>····················</td><td></td></tr>
<tr><td>J</td><td>6352</td><td>hCoV-19/England/ALDP-51DFB3/2020|EPI_ISL_559055|20</td><td>····················</td><td>····················</td><td>····················</td><td>····················</td><td>····················</td><td>····················</td><td></td></tr>
<tr><td>J</td><td>6353</td><td>hCoV-19/England/ALDP-523020/2020|EPI_ISL_559056|20</td><td>····················</td><td>····················</td><td>····················</td><td>····················</td><td>····················</td><td>····················</td><td></td></tr>
<tr><td>J</td><td>6354</td><td>hCoV-19/England/MILK-50ABF1/2020|EPI_ISL_559057|20</td><td>····················</td><td>····················</td><td>····················</td><td>····················</td><td>····················</td><td>····················</td><td></td></tr>
<tr><td>J</td><td>6355</td><td>hCoV-19/England/ALDP-52C072/2020|EPI_ISL_559059|20</td><td>····················</td><td>····················</td><td>····················</td><td>····················</td><td>····················</td><td>····················</td><td></td></tr>
<tr><td>J</td><td>6356</td><td>hCoV-19/Scotland/QEUH-99BF5F/2020|EPI_ISL_560000|2</td><td>····················</td><td>····················</td><td>····················</td><td>····················</td><td>····················</td><td>····················</td><td></td></tr>
<tr><td>J</td><td>6357</td><td>hCoV-19/Scotland/QEUH-99BF6E/2020|EPI_ISL_560001|2</td><td>····················</td><td>····················</td><td>····················</td><td>····················</td><td>····················</td><td>····················</td><td></td></tr>
<tr><td>J</td><td>6358</td><td>hCoV-19/Scotland/QEUH-99BF9B/2020|EPI_ISL_560002|2</td><td>····················</td><td>····················</td><td>····················</td><td>····················</td><td>····················</td><td>····················</td><td></td></tr>
<tr><td>J</td><td>6359</td><td>hCoV-19/Scotland/QEUH-99C370/2020|EPI_ISL_560003|2</td><td>····················</td><td>····················</td><td>····················</td><td>····················</td><td>····················</td><td>····················</td><td></td></tr>
<tr><td>J</td><td>6360</td><td>hCoV-19/Scotland/QEUH-9ACC45/2020|EPI_ISL_560004|2</td><td>····················</td><td>····················</td><td>····················</td><td>····················</td><td>····················</td><td>····················</td><td></td></tr>
<tr><td>J</td><td>6361</td><td>hCoV-19/Scotland/QEUH-9ACDAC/2020|EPI_ISL_560005|2</td><td>····················</td><td>····················</td><td>····················</td><td>····················</td><td>····················</td><td>····················</td><td></td></tr>
<tr><td>J</td><td>6362</td><td>hCoV-19/Scotland/QEUH-9ACDBB/2020|EPI_ISL_560006|2</td><td>····················</td><td>····················</td><td>····················</td><td>····················</td><td>····················</td><td>····················</td><td></td></tr>
<tr><td>J</td><td>6363</td><td>hCoV-19/Scotland/QEUH-9ACED6/2020|EPI_ISL_560007|2</td><td>····················</td><td>····················</td><td>····················</td><td>····················</td><td>····················</td><td>····················</td><td></td></tr>
<tr><td>J</td><td>6364</td><td>hCoV-19/Scotland/QEUH-9ACEE5/2020|EPI_ISL_560008|2</td><td>····················</td><td>····················</td><td>····················</td><td>····················</td><td>····················</td><td>····················</td><td></td></tr>
<tr><td>J</td><td>6365</td><td>hCoV-19/Scotland/QEUH-9ACEF4/2020|EPI_ISL_560009|2</td><td>····················</td><td>····················</td><td>····················</td><td>····················</td><td>····················</td><td>····················</td><td></td></tr>
<tr><td>J</td><td>6366</td><td>hCoV-19/Scotland/QEUH-9ACF2E/2020|EPI_ISL_560010|2</td><td>····················</td><td>····················</td><td>····················</td><td>····················</td><td>····················</td><td>····················</td><td></td></tr>
<tr><td>J</td><td>6367</td><td>hCoV-19/Scotland/QEUH-9ACF5B/2020|EPI_ISL_560011|2</td><td>····················</td><td>····················</td><td>····················</td><td>····················</td><td>····················</td><td>····················</td><td></td></tr>
<tr><td>J</td><td>6368</td><td>hCoV-19/Scotland/QEUH-9ACF97/2020|EPI_ISL_560012|2</td><td>····················</td><td>····················</td><td>····················</td><td>····················</td><td>····················</td><td>····················</td><td></td></tr>
<tr><td>J</td><td>6369</td><td>hCoV-19/Scotland/QEUH-9AD00C/2020|EPI_ISL_560013|2</td><td>····················</td><td>····················</td><td>····················</td><td>····················</td><td>····················</td><td>····················</td><td></td></tr>
<tr><td>J</td><td>6370</td><td>hCoV-19/Scotland/QEUH-9AD01B/2020|EPI_ISL_560014|2</td><td>····················</td><td>····················</td><td>····················</td><td>····················</td><td>····················</td><td>····················</td><td></td></tr>
<tr><td>J</td><td>6371</td><td>hCoV-19/Scotland/QEUH-9AD02A/2020|EPI_ISL_560015|2</td><td>····················</td><td>····················</td><td>····················</td><td>····················</td><td>····················</td><td>····················</td><td></td></tr>
<tr><td>J</td><td>6372</td><td>hCoV-19/Scotland/QEUH-9AD039/2020|EPI_ISL_560016|2</td><td>····················</td><td>····················</td><td>····················</td><td>····················</td><td>····················</td><td>····················</td><td></td></tr>
<tr><td>J</td><td>6373</td><td>hCoV-19/Scotland/QEUH-9AD066/2020|EPI_ISL_560017|2</td><td>····················</td><td>····················</td><td>····················</td><td>····················</td><td>····················</td><td>····················</td><td></td></tr>
<tr><td>J</td><td>6374</td><td>hCoV-19/Scotland/QEUH-9AD075/2020|EPI_ISL_560018|2</td><td>····················</td><td>····················</td><td>····················</td><td>····················</td><td>····················</td><td>····················</td><td></td></tr>
<tr><td>J</td><td>6375</td><td>hCoV-19/Scotland/QEUH-9AD0C0/2020|EPI_ISL_560019|2</td><td>····················</td><td>····················</td><td>····················</td><td>····················</td><td>····················</td><td>····················</td><td></td></tr>
<tr><td>J</td><td>6376</td><td>hCoV-19/Scotland/QEUH-9AD118/2020|EPI_ISL_560020|2</td><td>····················</td><td>····················</td><td>····················</td><td>····················</td><td>····················</td><td>····················</td><td></td></tr>
<tr><td>J</td><td>6377</td><td>hCoV-19/Scotland/QEUH-9AD1CD/2020|EPI_ISL_560021|2</td><td>····················</td><td>····················</td><td>····················</td><td>····················</td><td>····················</td><td>····················</td><td></td></tr>
<tr><td>J</td><td>6378</td><td>hCoV-19/Scotland/QEUH-9AD1DC/2020|EPI_ISL_560022|2</td><td>····················</td><td>····················</td><td>····················</td><td>····················</td><td>····················</td><td>····················</td><td></td></tr>
<tr><td>J</td><td>6379</td><td>hCoV-19/Scotland/QEUH-9AD29D/2020|EPI_ISL_560023|2</td><td>····················</td><td>····················</td><td>····················</td><td>····················</td><td>····················</td><td>····················</td><td></td></tr>
<tr><td>J</td><td>6380</td><td>hCoV-19/Scotland/QEUH-9AD2CA/2020|EPI_ISL_560024|2</td><td>····················</td><td>····················</td><td>····················</td><td>····················</td><td>····················</td><td>····················</td><td></td></tr>
<tr><td>J</td><td>6381</td><td>hCoV-19/Scotland/QEUH-9AF0CE/2020|EPI_ISL_560025|2</td><td>····················</td><td>····················</td><td>····················</td><td>····················</td><td>····················</td><td>····················</td><td></td></tr>
<tr><td>J</td><td>6382</td><td>hCoV-19/Scotland/QEUH-9AF0EC/2020|EPI_ISL_560026|2</td><td>····················</td><td>····················</td><td>····················</td><td>····················</td><td>····················</td><td>····················</td><td></td></tr>
<tr><td>J</td><td>6383</td><td>hCoV-19/Scotland/QEUH-9AF0FB/2020|EPI_ISL_560027|2</td><td>····················</td><td>····················</td><td>····················</td><td>····················</td><td>····················</td><td>····················</td><td></td></tr>
<tr><td>J</td><td>6384</td><td>hCoV-19/Scotland/QEUH-9AF107/2020|EPI_ISL_560028|2</td><td>····················</td><td>····················</td><td>····················</td><td>····················</td><td>····················</td><td>····················</td><td></td></tr>
<tr><td>J</td><td>6385</td><td>hCoV-19/Scotland/QEUH-9AF116/2020|EPI_ISL_560029|2</td><td>····················</td><td>····················</td><td>····················</td><td>····················</td><td>····················</td><td>····················</td><td></td></tr>
<tr><td>J</td><td>6386</td><td>hCoV-19/Scotland/QEUH-9AF143/2020|EPI_ISL_560030|2</td><td>····················</td><td>····················</td><td>····················</td><td>····················</td><td>····················</td><td>····················</td><td></td></tr>
<tr><td>J</td><td>6387</td><td>hCoV-19/Scotland/QEUH-9AF18F/2020|EPI_ISL_560031|2</td><td>····················</td><td>····················</td><td>····················</td><td>····················</td><td>····················</td><td>····················</td><td></td></tr>
<tr><td>J</td><td>6388</td><td>hCoV-19/Scotland/QEUH-9AF19E/2020|EPI_ISL_560032|2</td><td>····················</td><td>····················</td><td>····················</td><td>····················</td><td>····················</td><td>····················</td><td></td></tr>
<tr><td>J</td><td>6389</td><td>hCoV-19/Scotland/QEUH-9AF1CB/2020|EPI_ISL_560033|2</td><td>····················</td><td>····················</td><td>····················</td><td>····················</td><td>····················</td><td>····················</td><td></td></tr>
<tr><td>J</td><td>6390</td><td>hCoV-19/Scotland/QEUH-9AF3C5/2020|EPI_ISL_560034|2</td><td>····················</td><td>····················</td><td>····················</td><td>····················</td><td>····················</td><td>····················</td><td></td></tr>
<tr><td>J</td><td>6391</td><td>hCoV-19/Scotland/QEUH-9AF5CF/2020|EPI_ISL_560035|2</td><td>····················</td><td>····················</td><td>····················</td><td>····················</td><td>····················</td><td>····················</td><td></td></tr>
<tr><td>J</td><td>6392</td><td>hCoV-19/Scotland/QEUH-9B0BD8/2020|EPI_ISL_560036|2</td><td>····················</td><td>····················</td><td>····················</td><td>····················</td><td>····················</td><td>····················</td><td></td></tr>
<tr><td>J</td><td>6393</td><td>hCoV-19/Scotland/QEUH-9B0BE7/2020|EPI_ISL_560037|2</td><td>····················</td><td>····················</td><td>····················</td><td>····················</td><td>····················</td><td>····················</td><td></td></tr>
</table>

| | | | | | | | | |
|---|---|---|---|---|---|---|---|---|
| J | 6394 | hCoV-19/Scotland/QEUH-9B0BF6/2020\|EPI_ISL_560038\|2 | .................... | .................... | .................... | .................... | .................... | .................... |
| J | 6395 | hCoV-19/Scotland/QEUH-9B0C02/2020\|EPI_ISL_560039\|2 | .................... | .................... | .................... | .................... | .................... | .................... |
| J | 6396 | hCoV-19/Scotland/QEUH-9B0D4B/2020\|EPI_ISL_560040\|2 | .................... | .................... | .................... | .................... | .................... | .................... |
| J | 6397 | hCoV-19/Scotland/QEUH-9B0D78/2020\|EPI_ISL_560041\|2 | .................... | .................... | .................... | .................... | .................... | .................... |
| J | 6398 | hCoV-19/Scotland/QEUH-9B0D87/2020\|EPI_ISL_560042\|2 | .................... | .................... | ..T................. | .................... | .................... | .................... |
| J | 6399 | hCoV-19/Scotland/QEUH-9B0DC3/2020\|EPI_ISL_560043\|2 | .................... | .................... | .................... | .................... | .................... | .................... |
| J | 6400 | hCoV-19/Scotland/QEUH-9B0DD2/2020\|EPI_ISL_560044\|2 | .................... | .................... | .................... | .................... | .................... | .................... |
| J | 6401 | hCoV-19/England/NOTT-1134A3/2020\|EPI_ISL_549507\|20 | .................... | .A.................. | .................... | .................... | .................... | .................... |
| J | 6402 | hCoV-19/England/NOTT-1134D0/2020\|EPI_ISL_549508\|20 | .................... | .................... | .................... | .................... | .................... | .................... |
| J | 6403 | hCoV-19/England/NOTT-1134FE/2020\|EPI_ISL_549509\|20 | .................... | .................... | .................... | .................... | .................... | .................... |
| J | 6404 | hCoV-19/England/NOTT-11350A/2020\|EPI_ISL_549510\|20 | .................... | .A.................. | .................... | .................... | .................... | .................... |
| J | 6405 | hCoV-19/England/NOTT-113519/2020\|EPI_ISL_549511\|20 | .................... | .................... | .................... | .................... | .................... | .................... |
| J | 6406 | hCoV-19/England/NOTT-113555/2020\|EPI_ISL_549512\|20 | .................... | .................... | .................... | .................... | .................... | .................... |
| J | 6407 | hCoV-19/England/NOTT-113564/2020\|EPI_ISL_549513\|20 | .................... | .................... | .................... | .................... | .................... | .................... |
| J | 6408 | hCoV-19/England/NOTT-113573/2020\|EPI_ISL_549514\|20 | .................... | .................... | .................... | .................... | .................... | .................... |
| J | 6409 | hCoV-19/England/NOTT-113582/2020\|EPI_ISL_549515\|20 | .................... | .................... | .................... | .................... | .................... | .................... |
| J | 6410 | hCoV-19/England/NOTT-113591/2020\|EPI_ISL_549516\|20 | .................... | .................... | .................... | .................... | .................... | .................... |
| J | 6411 | hCoV-19/England/NOTT-1135A0/2020\|EPI_ISL_549517\|20 | .................... | .................... | .................... | .................... | ......T............. | .................... |
| J | 6412 | hCoV-19/England/NOTT-113616/2020\|EPI_ISL_549518\|20 | .................... | .................... | .................... | .................... | .................... | .................... |
| J | 6413 | hCoV-19/England/NOTT-113661/2020\|EPI_ISL_549519\|20 | .................... | .................... | .................... | .................... | .................... | .................... |
| J | 6414 | hCoV-19/England/NOTT-11369E/2020\|EPI_ISL_549520\|20 | .................... | .................... | .................... | .................... | .................... | .................... |
| J | 6415 | hCoV-19/England/NOTT-1136AD/2020\|EPI_ISL_549521\|20 | .................... | .................... | .................... | .................... | .................... | .................... |
| J | 6416 | hCoV-19/England/NOTT-1136CB/2020\|EPI_ISL_549522\|20 | .................... | .................... | .................... | .................... | .................... | .................... |
| J | 6417 | hCoV-19/England/QEUH-9962B5/2020\|EPI_ISL_549523\|20 | .................... | .................... | .................... | .................... | .................... | .................... |
| J | 6418 | hCoV-19/England/QEUH-9962F1/2020\|EPI_ISL_549524\|20 | .................... | .................... | .................... | .................... | .................... | .................... |
| J | 6419 | hCoV-19/England/QEUH-996349/2020\|EPI_ISL_549525\|20 | .................... | .................... | .................... | .................... | .................... | .................... |
| J | 6420 | hCoV-19/England/QEUH-996367/2020\|EPI_ISL_549526\|20 | .................... | .................... | .................... | .................... | .................... | .................... |
| J | 6421 | hCoV-19/England/QEUH-9963A3/2020\|EPI_ISL_549527\|20 | .................... | .................... | .................... | .................... | .................... | .................... |
| J | 6422 | hCoV-19/England/QEUH-9963B2/2020\|EPI_ISL_549528\|20 | .................... | .................... | .................... | .................... | .................... | .................... |
| J | 6423 | hCoV-19/England/QEUH-9963C1/2020\|EPI_ISL_549529\|20 | .................... | .................... | .................... | .................... | .................... | .................... |
| J | 6424 | hCoV-19/England/QEUH-9963D0/2020\|EPI_ISL_549530\|20 | .................... | .................... | .................... | .................... | .................... | .................... |
| J | 6425 | hCoV-19/England/QEUH-9963FE/2020\|EPI_ISL_549531\|20 | .................... | .................... | .................... | .................... | .................... | .................... |
| J | 6426 | hCoV-19/England/QEUH-996428/2020\|EPI_ISL_549532\|20 | .................... | .................... | .................... | .................... | .................... | .................... |
| J | 6427 | hCoV-19/England/QEUH-996482/2020\|EPI_ISL_549533\|20 | .................... | .................... | .................... | .................... | .................... | .................... |
| J | 6428 | hCoV-19/England/QEUH-9966B9/2020\|EPI_ISL_549534\|20 | .................... | .................... | .................... | .................... | .................... | .................... |
| J | 6429 | hCoV-19/England/QEUH-9966D7/2020\|EPI_ISL_549535\|20 | .................... | .................... | .................... | .................... | .................... | .................... |
| J | 6430 | hCoV-19/England/QEUH-99683B/2020\|EPI_ISL_549536\|20 | .................... | .................... | .................... | .................... | .................... | .................... |
| J | 6431 | hCoV-19/England/QEUH-996895/2020\|EPI_ISL_549537\|20 | .................... | .................... | .................... | .................... | .................... | .................... |
| J | 6432 | hCoV-19/England/QEUH-9968A4/2020\|EPI_ISL_549538\|20 | .................... | .................... | .................... | .................... | .................... | .................... |
| J | 6433 | hCoV-19/England/QEUH-9979CE/2020\|EPI_ISL_549539\|20 | .................... | .................... | .................... | .................... | .................... | .................... |
| J | 6434 | hCoV-19/England/MILK-977DE6/2020\|EPI_ISL_550500\|20 | .................... | .................... | .................... | .................... | .................... | .................... |
| J | 6435 | hCoV-19/England/MILK-978987/2020\|EPI_ISL_550503\|20 | .................... | .................... | .................... | .................... | .................... | .................... |
| J | 6436 | hCoV-19/England/MILK-977645/2020\|EPI_ISL_550504\|20 | .................... | .A.................. | .................... | .................... | .................... | .................... |
| J | 6437 | hCoV-19/England/MILK-978811/2020\|EPI_ISL_550505\|20 | .................... | .................... | .................... | .................... | .................... | .................... |
| J | 6438 | hCoV-19/England/MILK-97788B/2020\|EPI_ISL_550506\|20 | .................... | .................... | .................... | .................... | .................... | .................... |
| J | 6439 | hCoV-19/England/MILK-97787C/2020\|EPI_ISL_550507\|20 | .................... | .................... | .................... | .................... | .................... | .................... |
| J | 6440 | hCoV-19/England/MILK-97877E/2020\|EPI_ISL_550508\|20 | .................... | .................... | .................... | .................... | .................... | .................... |
| J | 6441 | hCoV-19/England/MILK-978644/2020\|EPI_ISL_550509\|20 | .................... | .................... | .................... | .................... | .................... | .................... |
| J | 6442 | hCoV-19/England/MILK-978662/2020\|EPI_ISL_550510\|20 | .................... | .................... | .................... | .................... | .................... | .................... |
| J | 6443 | hCoV-19/England/MILK-978A84/2020\|EPI_ISL_550511\|20 | .................... | .................... | .................... | .................... | .................... | .................... |
| J | 6444 | hCoV-19/England/MILK-978134/2020\|EPI_ISL_550514\|20 | .................... | .A.................. | .................... | .................... | .................... | ...A................ |
| J | 6445 | hCoV-19/England/MILK-9775FD/2020\|EPI_ISL_550515\|20 | .................... | .A.................. | .................... | .................... | .................... | ...A................ |
| J | 6446 | hCoV-19/England/MILK-977487/2020\|EPI_ISL_550516\|20 | .................... | .A.................. | .................... | .................... | .................... | ...A................ |
| J | 6447 | hCoV-19/England/MILK-99D913/2020\|EPI_ISL_550518\|20 | .................... | .................... | .................... | .................... | .................... | .................... |
| J | 6448 | hCoV-19/England/MILK-99E43F/2020\|EPI_ISL_550519\|20 | .................... | .................... | .................... | .................... | .................... | .................... |
| J | 6449 | hCoV-19/England/MILK-69E4C9/2020\|EPI_ISL_551813\|20 | .................... | .................... | .................... | .................... | .................... | .................... |
| J | 6450 | hCoV-19/England/MILK-6A625B/2020\|EPI_ISL_551820\|20 | .................... | .................... | .................... | .................... | .................... | .................... |
| J | 6451 | hCoV-19/England/ALDP-67FB6F/2020\|EPI_ISL_551825\|20 | .................... | .................... | .................... | .................... | .................... | .................... |
| J | 6452 | hCoV-19/England/ALDP-689652/2020\|EPI_ISL_551826\|20 | .................... | .................... | .................... | .................... | .................... | .................... |
| J | 6453 | hCoV-19/England/ALDP-688019/2020\|EPI_ISL_551827\|20 | .................... | .................... | .................... | .................... | .................... | .................... |
| J | 6454 | hCoV-19/England/MILK-6A831A/2020\|EPI_ISL_551828\|20 | .................... | .................... | .................... | .................... | .................... | .................... |
| J | 6455 | hCoV-19/England/MILK-69ECA3/2020\|EPI_ISL_551829\|20 | .................... | .................... | .................... | .................... | .................... | .................... |
| J | 6456 | hCoV-19/England/MILK-6A134E/2020\|EPI_ISL_551830\|20 | .................... | .................... | .................... | .................... | .................... | .................... |
| J | 6457 | hCoV-19/England/MILK-67A6DC/2020\|EPI_ISL_551831\|20 | .................... | .................... | .................... | .................... | .................... | .................... |
| J | 6458 | hCoV-19/England/ALDP-67D633/2020\|EPI_ISL_551832\|20 | .................... | .................... | .................... | .................... | .................... | .................... |
| J | 6459 | hCoV-19/England/MILK-699FEB/2020\|EPI_ISL_551833\|20 | .................... | .................... | .................... | .................... | .................... | .................... |
| J | 6460 | hCoV-19/England/ALDP-6832DC/2020\|EPI_ISL_551834\|20 | .................... | .................... | .................... | .................... | .................... | .................... |
| J | 6461 | hCoV-19/England/CAMC-991074/2020\|EPI_ISL_551835\|20 | .................... | .................... | .................... | .................... | ......T............. | .................... |
| J | 6462 | hCoV-19/England/CAMC-990B56/2020\|EPI_ISL_551836\|20 | .................... | .................... | .................... | .................... | .................... | .................... |
| J | 6463 | hCoV-19/England/CAMC-990DC9/2020\|EPI_ISL_551837\|20 | .................... | .................... | .................... | .................... | .................... | .................... |
| J | 6464 | hCoV-19/England/CAMC-990C53/2020\|EPI_ISL_551839\|20 | .................... | .................... | .................... | .................... | .................... | .................... |
| J | 6465 | hCoV-19/England/CAMC-990C35/2020\|EPI_ISL_551840\|20 | .................... | .................... | .................... | .................... | .................... | .................... |
| J | 6466 | hCoV-19/Wales/CAMC-991223/2020\|EPI_ISL_551841\|2020 | .................... | .................... | .................... | .................... | .................... | .................... |
| J | 6467 | hCoV-19/England/CAMC-990CAE/2020\|EPI_ISL_551842\|20 | .................... | .A.................. | .................... | .................... | .................... | ...A................ |
| J | 6468 | hCoV-19/England/CAMC-990EB7/2020\|EPI_ISL_551843\|20 | .................... | .................... | .................... | .................... | .................... | .................... |
| J | 6469 | hCoV-19/England/CAMC-990B92/2020\|EPI_ISL_551844\|20 | .................... | .................... | .................... | .................... | ......T............. | .................... |
| J | 6470 | hCoV-19/England/CAMC-990A95/2020\|EPI_ISL_551845\|20 | .................... | .................... | .................... | .................... | .................... | .................... |
| J | 6471 | hCoV-19/England/CAMC-9910FC/2020\|EPI_ISL_551846\|20 | .................... | .................... | .................... | .................... | .................... | .................... |
| J | 6472 | hCoV-19/England/CAMC-990B1A/2020\|EPI_ISL_551847\|20 | .................... | .................... | .................... | .................... | .................... | .................... |
| J | 6473 | hCoV-19/England/CAMC-99096B/2020\|EPI_ISL_551848\|20 | .................... | .A.................. | .................... | .................... | .................... | .................... |

| | | | | | | | | |
|---|---|---|---|---|---|---|---|---|
| J | 6474 | hCoV-19/England/ALDP-956442/2020\|EPI_ISL_554000\|20 | ···················· | ···················· | ····················· | ···················· | ···················· | ···················· |
| J | 6475 | hCoV-19/England/ALDP-955E09/2020\|EPI_ISL_554001\|20 | ···················· | ·A·················· | ····················· | ···················· | ···················· | ···················· |
| J | 6476 | hCoV-19/England/ALDP-9526D7/2020\|EPI_ISL_554006\|20 | ···················· | ···················· | ····················· | ···················· | ···················· | ···················· |
| J | 6477 | hCoV-19/England/ALDP-9563BE/2020\|EPI_ISL_554007\|20 | ···················· | ···················· | ····················· | ···················· | ···················· | ···················· |
| J | 6478 | hCoV-19/England/ALDP-95615A/2020\|EPI_ISL_554008\|20 | ···················· | ···················· | ····················· | ···················· | ···················· | ···················· |
| J | 6479 | hCoV-19/England/MILK-957696/2020\|EPI_ISL_554009\|20 | ···················· | ···················· | ····················· | ···················· | ···················· | ···················· |
| J | 6480 | hCoV-19/England/ALDP-955AD8/2020\|EPI_ISL_554010\|20 | ···················· | ···················· | ····················· | ···················· | ···················· | ···················· |
| J | 6481 | hCoV-19/England/ALDP-956178/2020\|EPI_ISL_554013\|20 | ···················· | ···················· | ····················· | ···················· | ···················· | ···················· |
| J | 6482 | hCoV-19/England/ALDP-9563AF/2020\|EPI_ISL_554014\|20 | ···················· | ···················· | ····················· | ···················· | ···················· | ···················· |
| J | 6483 | hCoV-19/England/MILK-957827/2020\|EPI_ISL_554015\|20 | ···················· | ···················· | ····················· | ···················· | ···················· | ···················· |
| J | 6484 | hCoV-19/England/ALDP-9560D5/2020\|EPI_ISL_554016\|20 | ···················· | ···················· | ····················· | ···················· | ···················· | ···················· |
| J | 6485 | hCoV-19/England/ALDP-95282C/2020\|EPI_ISL_554017\|20 | ···················· | ···················· | ····················· | ···················· | ···················· | ···················· |
| J | 6486 | hCoV-19/England/ALDP-955DA2/2020\|EPI_ISL_554018\|20 | ···················· | ···················· | ····················· | ···················· | ···················· | ···················· |
| J | 6487 | hCoV-19/England/ALDP-9562FD/2020\|EPI_ISL_554019\|20 | ···················· | ···················· | ····················· | ···················· | ···················· | ···················· |
| J | 6488 | hCoV-19/England/ALDP-95606C/2020\|EPI_ISL_554021\|20 | ···················· | ···················· | ····················· | ···················· | ···················· | ···················· |
| J | 6489 | hCoV-19/England/ALDP-95665B/2020\|EPI_ISL_554022\|20 | ···················· | ···················· | ····················· | ···················· | ···················· | ···················· |
| J | 6490 | hCoV-19/England/ALDP-95276B/2020\|EPI_ISL_554023\|20 | ···················· | ···················· | ····················· | ···················· | ···················· | ···················· |
| J | 6491 | hCoV-19/England/MILK-95755D/2020\|EPI_ISL_554024\|20 | ···················· | ···················· | ····················· | ···················· | ···················· | ···················· |
| J | 6492 | hCoV-19/England/MILK-9573CC/2020\|EPI_ISL_554025\|20 | ···················· | ···················· | ····················· | ···················· | ···················· | ···················· |
| J | 6493 | hCoV-19/England/MILK-9575E4/2020\|EPI_ISL_554026\|20 | ···················· | ···················· | ····················· | ···················· | ···················· | ···················· |
| J | 6494 | hCoV-19/England/ALDP-95656D/2020\|EPI_ISL_554027\|20 | ···················· | ···················· | ····················· | ···················· | ···················· | ···················· |
| J | 6495 | hCoV-19/England/MILK-957863/2020\|EPI_ISL_554028\|20 | ···················· | ·A·················· | ····················· | ···················· | ···················· | ···················· |
| J | 6496 | hCoV-19/England/ALDP-9560A8/2020\|EPI_ISL_554029\|20 | ···················· | ···················· | ····················· | ···················· | ···················· | ···················· |
| J | 6497 | hCoV-19/England/ALDP-9562C0/2020\|EPI_ISL_554030\|20 | ···················· | ···················· | ····················· | ···················· | ···················· | ···················· |
| J | 6498 | hCoV-19/England/ALDP-956169/2020\|EPI_ISL_554031\|20 | ···················· | ···················· | ····················· | ···················· | ···················· | ···················· |
| J | 6499 | hCoV-19/England/ALDP-955C78/2020\|EPI_ISL_554032\|20 | ···················· | ···················· | ····················· | ···················· | ···················· | ···················· |
| J | 6500 | hCoV-19/England/ALDP-955E54/2020\|EPI_ISL_554033\|20 | ···················· | ···················· | ····················· | ···················· | ···················· | ···················· |
| J | 6501 | hCoV-19/England/ALDP-952BE7/2020\|EPI_ISL_554035\|20 | ···················· | ···················· | ····················· | ···················· | ···················· | ···················· |
| J | 6502 | hCoV-19/England/ALDP-955B11/2020\|EPI_ISL_554036\|20 | ···················· | ···················· | ····················· | ···················· | ···················· | ···················· |
| J | 6503 | hCoV-19/England/ALDP-956451/2020\|EPI_ISL_554037\|20 | ···················· | ···················· | ····················· | ···················· | ···················· | ···················· |
| J | 6504 | hCoV-19/England/ALDP-95265F/2020\|EPI_ISL_554038\|20 | ···················· | ···················· | ····················· | ···················· | ···················· | ···················· |
| J | 6505 | hCoV-19/England/ALDP-955B7B/2020\|EPI_ISL_554044\|20 | ···················· | ···················· | ····················· | ···················· | ···················· | ···················· |
| J | 6506 | hCoV-19/England/ALDP-956257/2020\|EPI_ISL_554045\|20 | ···················· | ···················· | ····················· | ···················· | ···················· | ···················· |
| J | 6507 | hCoV-19/England/ALDP-955FF7/2020\|EPI_ISL_554047\|20 | ···················· | ···················· | ····················· | ···················· | ···················· | ···················· |
| J | 6508 | hCoV-19/England/MILK-9576A5/2020\|EPI_ISL_554048\|20 | ···················· | ···················· | ····················· | ···················· | ···················· | ···················· |
| J | 6509 | hCoV-19/England/ALDP-955D48/2020\|EPI_ISL_554049\|20 | ···················· | ···················· | ····················· | ···················· | ···················· | ···················· |
| J | 6510 | hCoV-19/England/ALDP-950123/2020\|EPI_ISL_555300\|20 | ···················· | ···················· | ····················· | ···················· | ···················· | ···················· |
| J | 6511 | hCoV-19/England/ALDP-94FD05/2020\|EPI_ISL_555301\|20 | ···················· | ···················· | ····················· | ···················· | ···················· | ···················· |
| J | 6512 | hCoV-19/England/ALDP-94FE99/2020\|EPI_ISL_555302\|20 | ···················· | ···················· | ····················· | ···················· | ···················· | ···················· |
| J | 6513 | hCoV-19/England/ALDP-94DF01/2020\|EPI_ISL_555305\|20 | ···················· | ···················· | ····················· | ···················· | ···················· | ···················· |
| J | 6514 | hCoV-19/England/ALDP-94E36E/2020\|EPI_ISL_555307\|20 | ···················· | ···················· | ····················· | ···················· | ···················· | ···················· |
| J | 6515 | hCoV-19/England/ALDP-94FBCF/2020\|EPI_ISL_555308\|20 | ···················· | ···················· | ····················· | ···················· | ···················· | ···················· |
| J | 6516 | hCoV-19/England/ALDP-94DA00/2020\|EPI_ISL_555309\|20 | ···················· | ···················· | ····················· | ···················· | ···················· | ···················· |
| J | 6517 | hCoV-19/England/ALDP-9502C6/2020\|EPI_ISL_555310\|20 | ···················· | ···················· | ····················· | ···················· | ···················· | ···················· |
| J | 6518 | hCoV-19/England/ALDP-950660/2020\|EPI_ISL_555311\|20 | ···················· | ···················· | ····················· | ···················· | ···················· | ···················· |
| J | 6519 | hCoV-19/England/ALDP-94E6CF/2020\|EPI_ISL_555312\|20 | ···················· | ···················· | ····················· | ···················· | ···················· | ···················· |
| J | 6520 | hCoV-19/England/ALDP-9519C0/2020\|EPI_ISL_555313\|20 | ···················· | ···················· | ····················· | ···················· | ···················· | ···················· |
| J | 6521 | hCoV-19/England/ALDP-95475A/2020\|EPI_ISL_555317\|20 | ···················· | ···················· | ····················· | ···················· | ···················· | ···················· |
| J | 6522 | hCoV-19/England/ALDP-9548FD/2020\|EPI_ISL_555318\|20 | ···················· | ···················· | ····················· | ···················· | ···················· | ···················· |
| J | 6523 | hCoV-19/England/ALDP-9518C3/2020\|EPI_ISL_555319\|20 | ···················· | ·A·················· | ····················· | ···················· | ···················· | ···················· |
| J | 6524 | hCoV-19/England/ALDP-952CB7/2020\|EPI_ISL_555320\|20 | ···················· | ···················· | ····················· | ···················· | ···················· | ···················· |
| J | 6525 | hCoV-19/England/ALDP-95480C/2020\|EPI_ISL_555321\|20 | ···················· | ···················· | ····················· | ···················· | ···················· | ···················· |
| J | 6526 | hCoV-19/England/ALDP-9548B1/2020\|EPI_ISL_555322\|20 | ···················· | ···················· | ····················· | ···················· | ···················· | ···················· |
| J | 6527 | hCoV-19/England/ALDP-94E0A3/2020\|EPI_ISL_555323\|20 | ···················· | ···················· | ····················· | ···················· | ···················· | ···················· |
| J | 6528 | hCoV-19/England/ALDP-954787/2020\|EPI_ISL_555324\|20 | ···················· | ···················· | ····················· | ···················· | ···················· | ···················· |
| J | 6529 | hCoV-19/England/ALDP-952B32/2020\|EPI_ISL_555325\|20 | ···················· | ···················· | ····················· | ···················· | ···················· | ···················· |
| J | 6530 | hCoV-19/England/ALDP-951C12/2020\|EPI_ISL_555327\|20 | ···················· | ···················· | ····················· | ···················· | ···················· | ···················· |
| J | 6531 | hCoV-19/England/ALDP-9518E1/2020\|EPI_ISL_555328\|20 | ···················· | ·A·················· | ····················· | ···················· | ···················· | ···················· |
| J | 6532 | hCoV-19/England/ALDP-955BE4/2020\|EPI_ISL_555329\|20 | ···················· | ···················· | ····················· | ···················· | ···················· | ···················· |
| J | 6533 | hCoV-19/England/ALDP-94E8AB/2020\|EPI_ISL_555330\|20 | ···················· | ···················· | ····················· | ···················· | ···················· | ···················· |
| J | 6534 | hCoV-19/England/ALDP-951C03/2020\|EPI_ISL_555333\|20 | ···················· | ···················· | ····················· | ···················· | ···················· | ···················· |
| J | 6535 | hCoV-19/England/ALDP-9559DB/2020\|EPI_ISL_555334\|20 | ···················· | ···················· | ····················· | ···················· | ···················· | ···················· |
| J | 6536 | hCoV-19/England/ALDP-9546B7/2020\|EPI_ISL_555335\|20 | ···················· | ···················· | ····················· | ···················· | ···················· | ···················· |
| J | 6537 | hCoV-19/England/ALDP-94E0B2/2020\|EPI_ISL_555336\|20 | ···················· | ···················· | ····················· | ···················· | ···················· | ···················· |
| J | 6538 | hCoV-19/England/ALDP-94E647/2020\|EPI_ISL_555339\|20 | ···················· | ···················· | ····················· | ···················· | ···················· | ···················· |
| J | 6539 | hCoV-19/England/ALDP-955A50/2020\|EPI_ISL_555340\|20 | ···················· | ···················· | ····················· | ···················· | ···················· | ···················· |
| J | 6540 | hCoV-19/England/ALDP-951869/2020\|EPI_ISL_555341\|20 | ···················· | ···················· | ····················· | ···················· | ···················· | ···················· |
| J | 6541 | hCoV-19/England/ALDP-9547D2/2020\|EPI_ISL_555342\|20 | ···················· | ···················· | ····················· | ···················· | ···················· | ···················· |
| J | 6542 | hCoV-19/England/ALDP-95182D/2020\|EPI_ISL_555343\|20 | ···················· | ···················· | ····················· | ···················· | ···················· | ···················· |
| J | 6543 | hCoV-19/England/ALDP-952BC9/2020\|EPI_ISL_555344\|20 | ···················· | ···················· | ····················· | ···················· | ···················· | ···················· |
| J | 6544 | hCoV-19/England/ALDP-9545C9/2020\|EPI_ISL_555345\|20 | ···················· | ···················· | ····················· | ···················· | ···················· | ···················· |
| J | 6545 | hCoV-19/England/ALDP-94E5C2/2020\|EPI_ISL_555346\|20 | ···················· | ···················· | ····················· | ···················· | ···················· | ···················· |
| J | 6546 | hCoV-19/England/ALDP-94E805/2020\|EPI_ISL_555347\|20 | ···················· | ···················· | ····················· | ···················· | ···················· | ···················· |
| J | 6547 | hCoV-19/England/ALDP-9546C6/2020\|EPI_ISL_555348\|20 | ···················· | ···················· | ····················· | ···················· | ···················· | ···················· |
| J | 6548 | hCoV-19/England/ALDP-954505/2020\|EPI_ISL_555349\|20 | ···················· | ···················· | ····················· | ···················· | ···················· | ···················· |
| J | 6549 | hCoV-19/England/ALDP-952BBA/2020\|EPI_ISL_555350\|20 | ···················· | ···················· | ····················· | ···················· | ···················· | ···················· |
| J | 6550 | hCoV-19/Wales/ALDP-952CA8/2020\|EPI_ISL_555351\|2020 | ···················· | ···················· | ····················· | ···················· | ···················· | ···················· |
| J | 6551 | hCoV-19/England/ALDP-952C02/2020\|EPI_ISL_555352\|20 | ···················· | ···················· | ····················· | ···················· | ···················· | ···················· |
| J | 6552 | hCoV-19/England/ALDP-94E5E0/2020\|EPI_ISL_555355\|20 | ···················· | ···················· | ····················· | ···················· | ···················· | ···················· |
| J | 6553 | hCoV-19/England/ALDP-95466C/2020\|EPI_ISL_555356\|20 | ···················· | ···················· | ····················· | ···················· | ···················· | ···················· |

| | | | | | | | | |
|---|---|---|---|---|---|---|---|---|
| J | 6554 | hCoV-19/England/ALDP-951B06/2020\|EPI_ISL_555358\|20 | .................... | .................... | ..................... | .................... | .................... | .................... |
| J | 6555 | hCoV-19/England/ALDP-94E674/2020\|EPI_ISL_555360\|20 | .................... | .................... | ..................... | .................... | .................... | .................... |
| J | 6556 | hCoV-19/England/ALDP-94E058/2020\|EPI_ISL_555361\|20 | .................... | .................... | ..................... | .................... | .................... | .................... |
| J | 6557 | hCoV-19/England/ALDP-955C3C/2020\|EPI_ISL_555363\|20 | .................... | .................... | ..................... | .................... | .................... | .................... |
| J | 6558 | hCoV-19/England/ALDP-954541/2020\|EPI_ISL_555364\|20 | .................... | .................... | ..................... | .................... | .................... | .................... |
| J | 6559 | hCoV-19/England/ALDP-95449F/2020\|EPI_ISL_555365\|20 | .................... | .................... | ..................... | .................... | .................... | .................... |
| J | 6560 | hCoV-19/England/ALDP-952B23/2020\|EPI_ISL_555366\|20 | .................... | .................... | ..................... | .................... | .................... | .................... |
| J | 6561 | hCoV-19/England/ALDP-95185A/2020\|EPI_ISL_555367\|20 | .................... | .................... | ..................... | .................... | .................... | .................... |
| J | 6562 | hCoV-19/England/ALDP-94E7F9/2020\|EPI_ISL_555375\|20 | .................... | .................... | ..................... | .................... | .................... | .................... |
| J | 6563 | hCoV-19/England/ALDP-9518A5/2020\|EPI_ISL_555376\|20 | .................... | .................... | ..................... | .................... | .................... | .................... |
| J | 6564 | hCoV-19/England/ALDP-955C2D/2020\|EPI_ISL_555378\|20 | .................... | .................... | ..................... | .................... | .................... | .................... |
| J | 6565 | hCoV-19/England/ALDP-94E61A/2020\|EPI_ISL_555379\|20 | .................... | .................... | ..................... | .................... | .................... | .................... |
| J | 6566 | hCoV-19/England/ALDP-8E4B45/2020\|EPI_ISL_556626\|20 | .................... | .A.................. | ..................... | .................... | .................... | .................... |
| J | 6567 | hCoV-19/England/ALDP-8BE379/2020\|EPI_ISL_556629\|20 | .................... | .................... | ..................... | .................... | .................... | .................... |
| J | 6568 | hCoV-19/England/ALDP-8F2670/2020\|EPI_ISL_556630\|20 | .................... | .................... | ..................... | .................... | .................... | .................... |
| J | 6569 | hCoV-19/England/ALDP-8C3E70/2020\|EPI_ISL_556631\|20 | .................... | .................... | ..................... | .................... | .................... | .................... |
| J | 6570 | hCoV-19/England/ALDP-8E5F48/2020\|EPI_ISL_556633\|20 | .................... | .................... | ..................... | .................... | .................... | .................... |
| J | 6571 | hCoV-19/England/ALDP-8E8EB1/2020\|EPI_ISL_556636\|20 | .................... | .................... | ..................... | .................... | .................... | .................... |
| J | 6572 | hCoV-19/England/CAMC-946050/2020\|EPI_ISL_556639\|20 | .................... | .................... | ..................... | .................... | .................... | .................... |
| J | 6573 | hCoV-19/England/CAMC-945D87/2020\|EPI_ISL_556640\|20 | .................... | .................... | ..................... | .................... | .................... | .................... |
| J | 6574 | hCoV-19/England/CAMC-94B02E/2020\|EPI_ISL_556641\|20 | .................... | .................... | ..................... | .................... | .................... | .................... |
| J | 6575 | hCoV-19/England/CAMC-9462B4/2020\|EPI_ISL_556642\|20 | .................... | .................... | ..................... | .................... | .................... | .................... |
| J | 6576 | hCoV-19/England/CAMC-946515/2020\|EPI_ISL_556643\|20 | .................... | .................... | ..................... | .................... | .................... | .................... |
| J | 6577 | hCoV-19/England/CAMC-94B71A/2020\|EPI_ISL_556646\|20 | .................... | .................... | ..................... | .................... | .................... | .................... |
| J | 6578 | hCoV-19/England/CAMC-94B228/2020\|EPI_ISL_556648\|20 | .................... | .................... | ..................... | .................... | .................... | .................... |
| J | 6579 | hCoV-19/England/CAMC-94C12A/2020\|EPI_ISL_556649\|20 | .................... | .................... | ..................... | .................... | .................... | .................... |
| J | 6580 | hCoV-19/England/CAMC-94C1FD/2020\|EPI_ISL_556650\|20 | .................... | .................... | ..................... | .................... | .................... | .................... |
| J | 6581 | hCoV-19/England/CAMC-945F36/2020\|EPI_ISL_556651\|20 | .................... | .................... | ..................... | .................... | .................... | .................... |
| J | 6582 | hCoV-19/England/CAMC-94A5D5/2020\|EPI_ISL_556654\|20 | .................... | .................... | ..................... | .................... | .................... | .................... |
| J | 6583 | hCoV-19/England/CAMC-946199/2020\|EPI_ISL_556655\|20 | .................... | .................... | ..................... | .................... | .................... | .................... |
| J | 6584 | hCoV-19/England/CAMC-94C333/2020\|EPI_ISL_556656\|20 | .................... | .................... | ..................... | .................... | .................... | .................... |
| J | 6585 | hCoV-19/England/CAMC-94BE51/2020\|EPI_ISL_556657\|20 | .................... | .................... | ..................... | .................... | .................... | .................... |
| J | 6586 | hCoV-19/England/CAMC-94665E/2020\|EPI_ISL_556658\|20 | .................... | ......A............. | ..................... | .................... | .................... | .................... |
| J | 6587 | hCoV-19/England/CAMC-94BEE8/2020\|EPI_ISL_556659\|20 | .................... | .................... | ..................... | .................... | .................... | .................... |
| J | 6588 | hCoV-19/England/CAMC-94B307/2020\|EPI_ISL_556660\|20 | .................... | .................... | ..................... | .................... | .................... | .................... |
| J | 6589 | hCoV-19/England/CAMC-94BD27/2020\|EPI_ISL_556661\|20 | .................... | .A.................. | ..................... | .................... | .................... | .................... |
| J | 6590 | hCoV-19/England/CAMC-9463C0/2020\|EPI_ISL_556662\|20 | .................... | .A.................. | ..................... | .................... | .................... | .................... |
| J | 6591 | hCoV-19/England/CAMC-94C3D9/2020\|EPI_ISL_556663\|20 | .................... | .................... | ..................... | .................... | .................... | .................... |
| J | 6592 | hCoV-19/England/CAMC-94B246/2020\|EPI_ISL_556665\|20 | .................... | .................... | ..................... | .................... | .................... | .................... |
| J | 6593 | hCoV-19/England/CAMC-94AD46/2020\|EPI_ISL_556666\|20 | .................... | .................... | ..................... | .................... | .................... | .................... |
| J | 6594 | hCoV-19/England/CAMC-94625A/2020\|EPI_ISL_556667\|20 | .................... | .................... | ..................... | .................... | .................... | .................... |
| J | 6595 | hCoV-19/England/CAMC-945C6C/2020\|EPI_ISL_556668\|20 | .................... | .A.................. | ..................... | .................... | .................... | .................... |
| J | 6596 | hCoV-19/England/CAMC-94C2CD/2020\|EPI_ISL_556669\|20 | .................... | .................... | ..................... | .................... | .................... | .................... |
| J | 6597 | hCoV-19/England/CAMC-946278/2020\|EPI_ISL_556670\|20 | .................... | .................... | ..................... | .................... | .................... | .................... |
| J | 6598 | hCoV-19/England/CAMC-945FCD/2020\|EPI_ISL_556671\|20 | .................... | .................... | ..................... | .................... | .................... | .................... |
| J | 6599 | hCoV-19/England/CAMC-9463EE/2020\|EPI_ISL_556672\|20 | .................... | ......A............. | ..................... | .................... | .................... | .................... |
| J | 6600 | hCoV-19/England/CAMC-9460BA/2020\|EPI_ISL_556673\|20 | .................... | .................... | ..................... | .................... | .................... | .................... |
| J | 6601 | hCoV-19/England/CAMC-945BD8/2020\|EPI_ISL_556674\|20 | .................... | .................... | ..................... | .................... | .................... | .................... |
| J | 6602 | hCoV-19/England/CAMC-946481/2020\|EPI_ISL_556675\|20 | .................... | .................... | ..................... | .................... | .................... | .................... |
| J | 6603 | hCoV-19/England/CAMC-94BE15/2020\|EPI_ISL_556676\|20 | .................... | .A.................. | ..................... | .................... | .................... | .................... |
| J | 6604 | hCoV-19/England/CAMC-94632A/2020\|EPI_ISL_556677\|20 | .................... | .................... | ..................... | .................... | .................... | .................... |
| J | 6605 | hCoV-19/England/CAMC-94B0F1/2020\|EPI_ISL_556678\|20 | .................... | .A.................. | ..................... | .................... | .................... | .................... |
| J | 6606 | hCoV-19/England/CAMC-945FBE/2020\|EPI_ISL_556679\|20 | .................... | .................... | ..................... | .................... | .................... | .................... |
| J | 6607 | hCoV-19/England/CAMC-9460D8/2020\|EPI_ISL_556681\|20 | .................... | .................... | ..................... | .................... | .................... | .................... |
| J | 6608 | hCoV-19/England/CAMC-94C209/2020\|EPI_ISL_556683\|20 | .................... | .................... | ..................... | .................... | .................... | .................... |
| J | 6609 | hCoV-19/England/CAMC-94C38E/2020\|EPI_ISL_556684\|20 | .................... | .................... | ..................... | .................... | .................... | .................... |
| J | 6610 | hCoV-19/England/CAMC-94C2FA/2020\|EPI_ISL_556685\|20 | .................... | .................... | ..................... | .................... | .................... | .................... |
| J | 6611 | hCoV-19/England/CAMC-94C2BE/2020\|EPI_ISL_556686\|20 | .................... | .............A...... | ..................... | .................... | .................... | .................... |
| J | 6612 | hCoV-19/England/CAMC-94C03C/2020\|EPI_ISL_556687\|20 | .................... | .................... | ..................... | .................... | .................... | .................... |
| J | 6613 | hCoV-19/England/CAMC-945F90/2020\|EPI_ISL_556688\|20 | .................... | .................... | ..................... | .................... | .................... | .................... |
| J | 6614 | hCoV-19/England/CAMC-946560/2020\|EPI_ISL_556689\|20 | .................... | .................... | ..................... | .................... | .................... | .................... |
| J | 6615 | hCoV-19/England/CAMC-94AAB8/2020\|EPI_ISL_556690\|20 | .................... | .................... | ..................... | .................... | .................... | .................... |
| J | 6616 | hCoV-19/England/CAMC-94BE60/2020\|EPI_ISL_556691\|20 | .................... | .A.................. | ..................... | .................... | .................... | .................... |
| J | 6617 | hCoV-19/England/CAMC-946551/2020\|EPI_ISL_556692\|20 | .................... | .................... | ..................... | .................... | .................... | .................... |
| J | 6618 | hCoV-19/England/CAMC-945BE7/2020\|EPI_ISL_556693\|20 | .................... | .................... | ..................... | .................... | .................... | .................... |
| J | 6619 | hCoV-19/England/CAMC-9461D5/2020\|EPI_ISL_556694\|20 | .................... | .................... | ..................... | .................... | .................... | .................... |
| J | 6620 | hCoV-19/England/CAMC-94C00F/2020\|EPI_ISL_556695\|20 | .................... | .................... | ..................... | .................... | .................... | .................... |
| J | 6621 | hCoV-19/England/CAMC-9464EB/2020\|EPI_ISL_556696\|20 | .................... | .................... | ..................... | .................... | .................... | .................... |
| J | 6622 | hCoV-19/England/CAMC-945CD5/2020\|EPI_ISL_556699\|20 | .................... | ......A............. | ..................... | .................... | .................... | .................... |
| J | 6623 | hCoV-19/England/ALDP-6BCC27/2020\|EPI_ISL_557955\|20 | .................... | .................... | ..................... | .................... | .................... | .................... |
| J | 6624 | hCoV-19/England/MILK-61AB75/2020\|EPI_ISL_557956\|20 | .................... | .................... | ..................... | .................... | .................... | .................... |
| J | 6625 | hCoV-19/England/MILK-61BEB7/2020\|EPI_ISL_557959\|20 | .................... | .................... | ...........T......... | .................... | .................... | .................... |
| J | 6626 | hCoV-19/England/ALDP-6B0072/2020\|EPI_ISL_557960\|20 | .................... | .................... | ..................... | .................... | .................... | .................... |
| J | 6627 | hCoV-19/England/ALDP-6BDEA8/2020\|EPI_ISL_557961\|20 | .................... | .................... | ..................... | .................... | .................... | .................... |
| J | 6628 | hCoV-19/Wales/ALDP-6AE7AF/2020\|EPI_ISL_557962\|2020 | .................... | .................... | ..................... | .................... | .................... | .................... |
| J | 6629 | hCoV-19/England/MILK-628628/2020\|EPI_ISL_557964\|20 | .................... | .................... | ..................... | .................... | .................... | .................... |
| J | 6630 | hCoV-19/England/MILK-620D49/2020\|EPI_ISL_557965\|20 | .................... | .................... | ..................... | .................... | .................... | .................... |
| J | 6631 | hCoV-19/England/ALDP-6B52FF/2020\|EPI_ISL_557968\|20 | .................... | .................... | ..................... | .................... | .................... | .................... |
| J | 6632 | hCoV-19/England/ALDP-6B656E/2020\|EPI_ISL_557971\|20 | .................... | .A.................. | ..................... | .................... | .................... | .................... |
| J | 6633 | hCoV-19/England/MILK-639CF5/2020\|EPI_ISL_557972\|20 | .................... | .................... | ..................... | .................... | .................... | .................... |

| | | | | | | | |
|---|---|---|---|---|---|---|---|
| J | 6634 hCoV-19/England/ALDP-6AEDF8/2020\|EPI_ISL_557973\|20 | ···················· | ···················· | ····················· | ···················· | ···················· | ···················· |
| J | 6635 hCoV-19/England/MILK-61C6BE/2020\|EPI_ISL_557974\|20 | ···················· | ···················· | ····················· | ···················· | ···················· | ···················· |
| J | 6636 hCoV-19/England/ALDP-6C270F/2020\|EPI_ISL_557975\|20 | ···················· | ···················· | ····················· | ···················· | ···················· | ···················· |
| J | 6637 hCoV-19/Wales/MILK-60FED4/2020\|EPI_ISL_557977\|2020 | ···················· | ···················· | ····················· | ···················· | ···················· | ···················· |
| J | 6638 hCoV-19/England/ALDP-6B712D/2020\|EPI_ISL_557978\|20 | ···················· | ···················· | ····················· | ···················· | ···················· | ···················· |
| J | 6639 hCoV-19/England/MILK-61C1DB/2020\|EPI_ISL_557979\|20 | ···················· | ···················· | ····················· | ···················· | ···················· | ···················· |
| J | 6640 hCoV-19/England/ALDP-6B35CB/2020\|EPI_ISL_557980\|20 | ···················· | ···················· | ····················· | ···················· | ···················· | ···················· |
| J | 6641 hCoV-19/England/ALDP-6C1B7D/2020\|EPI_ISL_557981\|20 | ···················· | ···················· | ····················· | ···················· | ···················· | ···················· |
| J | 6642 hCoV-19/England/ALDP-6AD667/2020\|EPI_ISL_557982\|20 | ···················· | ···················· | ····················· | ···················· | ···················· | ···················· |
| J | 6643 hCoV-19/England/ALDP-6AF1FB/2020\|EPI_ISL_557983\|20 | ···················· | ···················· | ····················· | ···················· | ···················· | ···················· |
| J | 6644 hCoV-19/England/MILK-622EBD/2020\|EPI_ISL_557984\|20 | ···················· | ···················· | ····················· | ···················· | ···················· | ···················· |
| J | 6645 hCoV-19/England/ALDP-6BE8D6/2020\|EPI_ISL_557985\|20 | ···················· | ···················· | ····················· | ···················· | ···················· | ···················· |
| J | 6646 hCoV-19/England/ALDP-658A7E/2020\|EPI_ISL_557986\|20 | ···················· | ···················· | ····················· | ···················· | ···················· | ···················· |
| J | 6647 hCoV-19/England/ALDP-68FF8E/2020\|EPI_ISL_557987\|20 | ···················· | ···················· | ····················· | ···················· | ···················· | ···················· |
| J | 6648 hCoV-19/England/MILK-6555AD/2020\|EPI_ISL_557989\|20 | ···················· | ···················· | ····················· | ···················· | ···················· | ···················· |
| J | 6649 hCoV-19/England/ALDP-651C9D/2020\|EPI_ISL_557990\|20 | ···················· | ···················· | ····················· | ···················· | ···················· | ···················· |
| J | 6650 hCoV-19/England/ALDP-6762FA/2020\|EPI_ISL_557991\|20 | ···················· | ···················· | ····················· | ···················· | ···················· | ···················· |
| J | 6651 hCoV-19/England/ALDP-69343D/2020\|EPI_ISL_557992\|20 | ···················· | ···················· | ····················· | ···················· | ···················· | ···················· |
| J | 6652 hCoV-19/England/ALDP-66AC0A/2020\|EPI_ISL_557993\|20 | ···················· | ···················· | ····················· | ···················· | ···················· | ···················· |
| J | 6653 hCoV-19/England/ALDP-6456CB/2020\|EPI_ISL_557994\|20 | ···················· | ···················· | ····················· | ···················· | ···················· | ···················· |
| J | 6654 hCoV-19/England/ALDP-67361F/2020\|EPI_ISL_557995\|20 | ···················· | ···················· | ····················· | ···················· | ···················· | ···················· |
| J | 6655 hCoV-19/Wales/ALDP-68A28A/2020\|EPI_ISL_557996\|2020 | ···················· | ···················· | ····················· | ···················· | ···················· | ···················· |
| J | 6656 hCoV-19/England/ALDP-64B601/2020\|EPI_ISL_557998\|20 | ···················· | ···················· | ····················· | ···················· | ···················· | ···················· |
| J | 6657 hCoV-19/United Arab Emirates/DU-A-S3-3_S9/2020\|EPI | ···················· | ···················· | ····················· | ···················· | ···················· | ···················· |
| J | 6658 hCoV-19/United Arab Emirates/DU-A-S9-2_S4/2020\|EPI | ···················· | ···················· | ····················· | ···················· | ···················· | ···················· |
| J | 6659 hCoV-19/United Arab Emirates/DU-A-20-205_S1/2020\|E | ···················· | ···················· | ····················· | ···················· | ···················· | ···················· |
| J | 6660 hCoV-19/Singapore/963/2020\|EPI_ISL_548972\|2020-09- | ···················· | ···················· | ····················· | ···················· | ···················· | ···················· |
| J | 6661 hCoV-19/Singapore/964/2020\|EPI_ISL_548973\|2020-09- | ···················· | ···················· | ····················· | ···················· | ···················· | ···················· |
| J | 6662 hCoV-19/Singapore/965/2020\|EPI_ISL_548974\|2020-09- | ···················· | ···················· | ····················· | ···················· | ···················· | ···················· |
| J | 6663 hCoV-19/Singapore/969/2020\|EPI_ISL_548977\|2020-03- | ···················· | ···················· | ····················· | ···················· | ···················· | ···················· |
| J | 6664 hCoV-19/Singapore/970/2020\|EPI_ISL_548978\|2020-03- | ···················· | ···················· | ····················· | ···················· | ···················· | ···················· |
| J | 6665 hCoV-19/Singapore/972/2020\|EPI_ISL_548980\|2020-03- | ···················· | ···················· | ····················· | ···················· | ···················· | ···················· |
| J | 6666 hCoV-19/Singapore/973/2020\|EPI_ISL_548981\|2020-03- | ···················· | ···················· | ····················· | ···················· | ···················· | ···················· |
| J | 6667 hCoV-19/Singapore/974/2020\|EPI_ISL_548982\|2020-03- | ···················· | ···················· | ····················· | ···················· | ···················· | ···················· |
| J | 6668 hCoV-19/Singapore/975/2020\|EPI_ISL_548983\|2020-03- | ···················· | ···················· | ····················· | ···················· | ···················· | ···················· |
| J | 6669 hCoV-19/Singapore/976/2020\|EPI_ISL_548984\|2020-03- | ···················· | ···················· | ····················· | ···················· | ···················· | ···················· |
| J | 6670 hCoV-19/Singapore/977/2020\|EPI_ISL_548985\|2020-03- | ···················· | ···················· | ····················· | ···················· | ···················· | ···················· |
| J | 6671 hCoV-19/England/QEUH-997A61/2020\|EPI_ISL_549540\|20 | ···················· | ···················· | ····················· | ···················· | ····T··············· | ···················· |
| J | 6672 hCoV-19/England/QEUH-997A70/2020\|EPI_ISL_549541\|20 | ···················· | ···················· | ····················· | ···················· | ···················· | ···················· |
| J | 6673 hCoV-19/England/QEUH-997A8F/2020\|EPI_ISL_549542\|20 | ···················· | ···················· | ····················· | ···················· | ···················· | ···················· |
| J | 6674 hCoV-19/England/QEUH-997BAA/2020\|EPI_ISL_549543\|20 | ···················· | ···················· | ····················· | ···················· | ···················· | ···················· |
| J | 6675 hCoV-19/England/QEUH-997BB9/2020\|EPI_ISL_549544\|20 | ···················· | ···················· | ····················· | ···················· | ···················· | ···················· |
| J | 6676 hCoV-19/England/QEUH-997C01/2020\|EPI_ISL_549545\|20 | ···················· | ···················· | ····················· | ···················· | ···················· | ···················· |
| J | 6677 hCoV-19/England/QEUH-997C10/2020\|EPI_ISL_549546\|20 | ···················· | ···················· | ····················· | ···················· | ···················· | ···················· |
| J | 6678 hCoV-19/England/QEUH-9B9119/2020\|EPI_ISL_549547\|20 | ···················· | ···················· | ····················· | ···················· | ···················· | ···················· |
| J | 6679 hCoV-19/England/QEUH-9B9216/2020\|EPI_ISL_549548\|20 | ···················· | ···················· | ····················· | ···················· | ···················· | ···················· |
| J | 6680 hCoV-19/England/QEUH-9B9A4B/2020\|EPI_ISL_549549\|20 | ···················· | ···················· | ····················· | ···················· | ···················· | ···················· |
| J | 6681 hCoV-19/England/QEUH-9B8BFE/2020\|EPI_ISL_549551\|20 | ···················· | ···················· | ····················· | ···················· | ···················· | ···················· |
| J | 6682 hCoV-19/England/QEUH-9B8FA7/2020\|EPI_ISL_549552\|20 | ···················· | ···················· | ····················· | ···················· | ······T············· | ···················· |
| J | 6683 hCoV-19/England/QEUH-9B93F5/2020\|EPI_ISL_549553\|20 | ···················· | ···················· | ····················· | ···················· | ···················· | ···················· |
| J | 6684 hCoV-19/England/QEUH-9B9C54/2020\|EPI_ISL_549554\|20 | ···················· | ···················· | ····················· | ···················· | ···················· | ···················· |
| J | 6685 hCoV-19/England/QEUH-9B8578/2020\|EPI_ISL_549555\|20 | ···················· | ···················· | ····················· | ···················· | ···················· | ···················· |
| J | 6686 hCoV-19/England/QEUH-9B9CBE/2020\|EPI_ISL_549556\|20 | ···················· | ···················· | ····················· | ···················· | ···················· | ···················· |
| J | 6687 hCoV-19/England/QEUH-9B8BA3/2020\|EPI_ISL_549557\|20 | ···················· | ···················· | ····················· | ···················· | ···················· | ···················· |
| J | 6688 hCoV-19/England/QEUH-9B9234/2020\|EPI_ISL_549558\|20 | ···················· | ···················· | ····················· | ···················· | ···················· | ···················· |
| J | 6689 hCoV-19/England/QEUH-9B9C90/2020\|EPI_ISL_549559\|20 | ···················· | ·A·················· | ····················· | ···················· | ···················· | ···A················ |
| J | 6690 hCoV-19/England/QEUH-9B9CEB/2020\|EPI_ISL_549560\|20 | ···················· | ···················· | ····················· | ···················· | ···················· | ···················· |
| J | 6691 hCoV-19/England/QEUH-9B937D/2020\|EPI_ISL_549561\|20 | ···················· | ···················· | ····················· | ···················· | ···················· | ···················· |
| J | 6692 hCoV-19/England/QEUH-9B91A0/2020\|EPI_ISL_549562\|20 | ···················· | ···················· | ····················· | ···················· | ···················· | ···················· |
| J | 6693 hCoV-19/England/QEUH-9B8DE9/2020\|EPI_ISL_549563\|20 | ···················· | ···················· | ····················· | ···················· | ···················· | ···················· |
| J | 6694 hCoV-19/England/QEUH-9B86A2/2020\|EPI_ISL_549566\|20 | ···················· | ···················· | ····················· | ···················· | ···················· | ···················· |
| J | 6695 hCoV-19/England/QEUH-9B843F/2020\|EPI_ISL_549567\|20 | ···················· | ···················· | ····················· | ···················· | ···················· | ···················· |
| J | 6696 hCoV-19/England/QEUH-9B8790/2020\|EPI_ISL_549568\|20 | ···················· | ···················· | ····················· | ···················· | ···················· | ···················· |
| J | 6697 hCoV-19/England/QEUH-9B9A0F/2020\|EPI_ISL_549569\|20 | ···················· | ···················· | ····················· | ···················· | ···················· | ···················· |
| J | 6698 hCoV-19/England/QEUH-9B9AD2/2020\|EPI_ISL_549570\|20 | ···················· | ···················· | ····················· | ···················· | ···················· | ···················· |
| J | 6699 hCoV-19/England/QEUH-9B9C18/2020\|EPI_ISL_549571\|20 | ···················· | ···················· | ····················· | ···················· | ···················· | ···················· |
| J | 6700 hCoV-19/England/QEUH-9B9137/2020\|EPI_ISL_549573\|20 | ···················· | ···················· | ····················· | ···················· | ···················· | ···················· |
| J | 6701 hCoV-19/England/QEUH-9B8B76/2020\|EPI_ISL_549574\|20 | ···················· | ·A·······A·········· | ····················· | ···················· | ···················· | ···················· |
| J | 6702 hCoV-19/England/QEUH-9B8E40/2020\|EPI_ISL_549575\|20 | ···················· | ···················· | ····················· | ···················· | ···················· | ···················· |
| J | 6703 hCoV-19/England/QEUH-9B86C0/2020\|EPI_ISL_549576\|20 | ···················· | ···················· | ····················· | ···················· | ····T··············· | ···················· |
| J | 6704 hCoV-19/England/QEUH-9B88D9/2020\|EPI_ISL_549577\|20 | ···················· | ···················· | ····················· | ···················· | ···················· | ···················· |
| J | 6705 hCoV-19/England/QEUH-9B88E8/2020\|EPI_ISL_549578\|20 | ···················· | ···················· | ····················· | ···················· | ···················· | ···················· |
| J | 6706 hCoV-19/England/QEUH-9B9058/2020\|EPI_ISL_549579\|20 | ···················· | ···················· | ····················· | ···················· | ···················· | ···················· |
| J | 6707 hCoV-19/England/QEUH-9B9C45/2020\|EPI_ISL_549581\|20 | ···················· | ···················· | ····················· | ···················· | ···················· | ···················· |
| J | 6708 hCoV-19/England/QEUH-9B89E5/2020\|EPI_ISL_549582\|20 | ···················· | ·A·················· | ····················· | ···················· | ···················· | ···················· |
| J | 6709 hCoV-19/England/QEUH-9B8F4D/2020\|EPI_ISL_549583\|20 | ···················· | ···················· | ····················· | ···················· | ···················· | ···················· |
| J | 6710 hCoV-19/England/QEUH-9B8CBF/2020\|EPI_ISL_549585\|20 | ···················· | ···················· | ····················· | ···················· | ···················· | ···················· |
| J | 6711 hCoV-19/England/QEUH-9B87BE/2020\|EPI_ISL_549586\|20 | ···················· | ···················· | ····················· | ···················· | ···················· | ···················· |
| J | 6712 hCoV-19/England/QEUH-9B8C19/2020\|EPI_ISL_549587\|20 | ···················· | ···················· | ····················· | ···················· | ···················· | ···················· |
| J | 6713 hCoV-19/England/QEUH-9B9E21/2020\|EPI_ISL_549588\|20 | ···················· | ···················· | ····················· | ···················· | ···················· | ···················· |

| | | | | | | | |
|---|---|---|---|---|---|---|---|
| J | 6714 hCoV-19/England/QEUH-9B8350/2020\|EPI_ISL_549589\|20 | ···················· | ···················· | ···················· | ···················· | ···················· | ···················· |
| J | 6715 hCoV-19/England/QEUH-9B8BD0/2020\|EPI_ISL_549591\|20 | ···················· | ···················· | ···················· | ···················· | ···················· | ···················· |
| J | 6716 hCoV-19/England/QEUH-9B92BC/2020\|EPI_ISL_549593\|20 | ···················· | ···················· | ···················· | ···················· | ···················· | ···················· |
| J | 6717 hCoV-19/England/QEUH-9B85C3/2020\|EPI_ISL_549595\|20 | ···················· | ···················· | ···················· | ···················· | ···················· | ···················· |
| J | 6718 hCoV-19/England/QEUH-9B9D06/2020\|EPI_ISL_549596\|20 | ···················· | ···················· | ···················· | ···················· | ···················· | ···················· |
| J | 6719 hCoV-19/England/QEUH-9B895E/2020\|EPI_ISL_549597\|20 | ···················· | ·A·················· | ···················· | ···················· | ···················· | ··A················· |
| J | 6720 hCoV-19/England/QEUH-9B8772/2020\|EPI_ISL_549598\|20 | ···················· | ···················· | ···················· | ···················· | ·····T·············· | ···················· |
| J | 6721 hCoV-19/England/QEUH-9B83AB/2020\|EPI_ISL_549599\|20 | ···················· | ···················· | ···················· | ···················· | ···················· | ···················· |
| J | 6722 hCoV-19/England/MILK-99DE7E/2020\|EPI_ISL_550521\|20 | ···················· | ···················· | ···················· | ···················· | ···················· | ···················· |
| J | 6723 hCoV-19/England/MILK-99DCFC/2020\|EPI_ISL_550522\|20 | ···················· | ···················· | ···················· | ···················· | ···················· | ···················· |
| J | 6724 hCoV-19/England/MILK-99D89E/2020\|EPI_ISL_550523\|20 | ···················· | ···················· | ···················· | ···················· | ···················· | ···················· |
| J | 6725 hCoV-19/England/MILK-99E3BA/2020\|EPI_ISL_550524\|20 | ···················· | ···················· | ···················· | ···················· | ···················· | ···················· |
| J | 6726 hCoV-19/England/MILK-99DDCC/2020\|EPI_ISL_550525\|20 | ···················· | ········A··········· | ···················· | ···················· | ···················· | ···················· |
| J | 6727 hCoV-19/England/MILK-99E2BD/2020\|EPI_ISL_550528\|20 | ···················· | ···················· | ···················· | ···················· | ···················· | ···················· |
| J | 6728 hCoV-19/England/MILK-99C501/2020\|EPI_ISL_550529\|20 | ···················· | ···················· | ···················· | ···················· | ···················· | ···················· |
| J | 6729 hCoV-19/England/MILK-99D95F/2020\|EPI_ISL_550530\|20 | ···················· | ···················· | ···················· | ···················· | ···················· | ···················· |
| J | 6730 hCoV-19/England/MILK-99E077/2020\|EPI_ISL_550531\|20 | ···················· | ···················· | ···················· | ···················· | ···················· | ···················· |
| J | 6731 hCoV-19/England/MILK-99E129/2020\|EPI_ISL_550532\|20 | ···················· | ···················· | ··········T········· | ···················· | ···················· | ···················· |
| J | 6732 hCoV-19/England/MILK-99DDBD/2020\|EPI_ISL_550533\|20 | ···················· | ···················· | ···················· | ···················· | ···················· | ···················· |
| J | 6733 hCoV-19/England/MILK-99E095/2020\|EPI_ISL_550534\|20 | ···················· | ···················· | ···················· | ···················· | ···················· | ···················· |
| J | 6734 hCoV-19/Wales/MILK-99C54D/2020\|EPI_ISL_550535\|2020 | ···················· | ···················· | ···················· | ···················· | ···················· | ···················· |
| J | 6735 hCoV-19/Wales/MILK-99DA5C/2020\|EPI_ISL_550536\|2020 | ···················· | ···················· | ···················· | ···················· | ···················· | ···················· |
| J | 6736 hCoV-19/England/MILK-99C413/2020\|EPI_ISL_550538\|20 | ···················· | ·····A·············· | ···················· | ···················· | ···················· | ···················· |
| J | 6737 hCoV-19/England/MILK-99E3E7/2020\|EPI_ISL_550539\|20 | ···················· | ···················· | ···················· | ···················· | ···················· | ···················· |
| J | 6738 hCoV-19/England/MILK-99E10B/2020\|EPI_ISL_550540\|20 | ···················· | ···················· | ···················· | ···················· | ···················· | ···················· |
| J | 6739 hCoV-19/England/MILK-99DBD1/2020\|EPI_ISL_550541\|20 | ···················· | ·A·················· | ···················· | ···················· | ···················· | ···················· |
| J | 6741 hCoV-19/England/MILK-99DF4E/2020\|EPI_ISL_550543\|20 | ···················· | ···················· | ···················· | ···················· | ···················· | ···················· |
| J | 6742 hCoV-19/England/MILK-99DFB7/2020\|EPI_ISL_550544\|20 | ···················· | ·A·················· | ···················· | ···················· | ···················· | ··A················· |
| J | 6743 hCoV-19/England/MILK-99E314/2020\|EPI_ISL_550545\|20 | ···················· | ···················· | ···················· | ···················· | ···················· | ···················· |
| J | 6744 hCoV-19/England/MILK-99E1B0/2020\|EPI_ISL_550547\|20 | ···················· | ···················· | ···················· | ···················· | ···················· | ···················· |
| J | 6745 hCoV-19/England/MILK-99DCCF/2020\|EPI_ISL_550548\|20 | ···················· | ···················· | ···················· | ···················· | ···················· | ···················· |
| J | 6746 hCoV-19/England/MILK-99DEBA/2020\|EPI_ISL_550549\|20 | ···················· | ···················· | ···················· | ···················· | ···················· | ···················· |
| J | 6747 hCoV-19/England/MILK-99E3AB/2020\|EPI_ISL_550550\|20 | ···················· | ···················· | ···················· | ···················· | ···················· | ···················· |
| J | 6748 hCoV-19/Wales/MILK-99C9D8/2020\|EPI_ISL_550551\|2020 | ···················· | ···················· | ···················· | ···················· | ···················· | ···················· |
| J | 6749 hCoV-19/Wales/MILK-99C52F/2020\|EPI_ISL_550552\|2020 | ···················· | ···················· | ···················· | ···················· | ···················· | ···················· |
| J | 6750 hCoV-19/England/MILK-99E341/2020\|EPI_ISL_550553\|20 | ···················· | ···················· | ···················· | ···················· | ···················· | ···················· |
| J | 6751 hCoV-19/England/MILK-99C668/2020\|EPI_ISL_550554\|20 | ···················· | ···················· | ···················· | ···················· | ···················· | ···················· |
| J | 6752 hCoV-19/England/MILK-99C844/2020\|EPI_ISL_550555\|20 | ···················· | ···················· | ···················· | ···················· | ···················· | ···················· |
| J | 6753 hCoV-19/England/MILK-99C8DB/2020\|EPI_ISL_550556\|20 | ···················· | ···················· | ···················· | ···················· | ···················· | ···················· |
| J | 6754 hCoV-19/England/MILK-99D157/2020\|EPI_ISL_550557\|20 | ···················· | ···················· | ···················· | ···················· | ···················· | ···················· |
| J | 6755 hCoV-19/England/MILK-99E147/2020\|EPI_ISL_550558\|20 | ···················· | ···················· | ···················· | ···················· | ···················· | ···················· |
| J | 6756 hCoV-19/England/MILK-99E271/2020\|EPI_ISL_550560\|20 | ···················· | ···················· | ···················· | ···················· | ···················· | ···················· |
| J | 6757 hCoV-19/England/MILK-99C8EA/2020\|EPI_ISL_550561\|20 | ···················· | ···················· | ···················· | ···················· | ···················· | ···················· |
| J | 6758 hCoV-19/England/MILK-99DB3B/2020\|EPI_ISL_550563\|20 | ···················· | ···················· | ···················· | ···················· | ···················· | ···················· |
| J | 6759 hCoV-19/England/MILK-99E305/2020\|EPI_ISL_550564\|20 | ···················· | ···················· | ···················· | ···················· | ·····T·············· | ···················· |
| J | 6760 hCoV-19/England/MILK-99C62C/2020\|EPI_ISL_550566\|20 | ···················· | ···················· | ···················· | ···················· | ···················· | ···················· |
| J | 6761 hCoV-19/England/MILK-99CB1E/2020\|EPI_ISL_550567\|20 | ···················· | ···················· | ···················· | ···················· | ···················· | ···················· |
| J | 6762 hCoV-19/England/MILK-99DAA7/2020\|EPI_ISL_550568\|20 | ···················· | ···················· | ···················· | ···················· | ···················· | ···················· |
| J | 6763 hCoV-19/England/MILK-99C8AE/2020\|EPI_ISL_550569\|20 | ···················· | ···················· | ···················· | ···················· | ···················· | ···················· |
| J | 6764 hCoV-19/England/MILK-99E53C/2020\|EPI_ISL_550570\|20 | ···················· | ···················· | ···················· | ···················· | ···················· | ···················· |
| J | 6765 hCoV-19/England/MILK-99C6B3/2020\|EPI_ISL_550571\|20 | ···················· | ···················· | ···················· | ···················· | ···················· | ···················· |
| J | 6766 hCoV-19/England/MILK-99C4E6/2020\|EPI_ISL_550572\|20 | ···················· | ···················· | ···················· | ···················· | ···················· | ···················· |
| J | 6767 hCoV-19/England/MILK-99CA02/2020\|EPI_ISL_550573\|20 | ···················· | ·A·················· | ···················· | ···················· | ···················· | ··A················· |
| J | 6768 hCoV-19/England/MILK-99D8F8/2020\|EPI_ISL_550574\|20 | ···················· | ···················· | ···················· | ···················· | ···················· | ···················· |
| J | 6769 hCoV-19/Wales/MILK-99C9C9/2020\|EPI_ISL_550575\|2020 | ···················· | ···················· | ···················· | ···················· | ···················· | ···················· |
| J | 6770 hCoV-19/England/MILK-99E4C6/2020\|EPI_ISL_550576\|20 | ···················· | ···················· | ···················· | ···················· | ···················· | ···················· |
| J | 6771 hCoV-19/England/MILK-99C5E3/2020\|EPI_ISL_550577\|20 | ···················· | ···················· | ···················· | ···················· | ···················· | ···················· |
| J | 6772 hCoV-19/England/MILK-99CAE4/2020\|EPI_ISL_550578\|20 | ···················· | ···················· | ···················· | ···················· | ···················· | ···················· |
| J | 6773 hCoV-19/England/MILK-99E332/2020\|EPI_ISL_550579\|20 | ···················· | ···················· | ···················· | ···················· | ···················· | ···················· |
| J | 6774 hCoV-19/England/MILK-99DA2F/2020\|EPI_ISL_550580\|20 | ···················· | ···················· | ···················· | ···················· | ···················· | ···················· |
| J | 6775 hCoV-19/England/MILK-99C4D7/2020\|EPI_ISL_550582\|20 | ···················· | ···················· | ···················· | ···················· | ···················· | ···················· |
| J | 6776 hCoV-19/England/MILK-99C6FF/2020\|EPI_ISL_550583\|20 | ···················· | ···················· | ···················· | ···················· | ·····T·············· | ···················· |
| J | 6777 hCoV-19/England/MILK-99C659/2020\|EPI_ISL_550584\|20 | ···················· | ···················· | ···················· | ···················· | ···················· | ···················· |
| J | 6778 hCoV-19/England/MILK-99DBE0/2020\|EPI_ISL_550585\|20 | ···················· | ···················· | ···················· | ···················· | ···················· | ···················· |
| J | 6779 hCoV-19/England/MILK-99C6A4/2020\|EPI_ISL_550586\|20 | ···················· | ···················· | ···················· | ···················· | ···················· | ···················· |
| J | 6780 hCoV-19/Wales/MILK-99C7DE/2020\|EPI_ISL_550587\|2020 | ···················· | ·A·················· | ···················· | ···················· | ···················· | ··A················· |
| J | 6781 hCoV-19/England/MILK-99DB2C/2020\|EPI_ISL_550588\|20 | ···················· | ···················· | ···················· | ···················· | ···················· | ···················· |
| J | 6782 hCoV-19/England/MILK-99D7CE/2020\|EPI_ISL_550589\|20 | ···················· | ···················· | ···················· | ···················· | ···················· | ···················· |
| J | 6783 hCoV-19/England/MILK-99CA11/2020\|EPI_ISL_550591\|20 | ···················· | ···················· | ···················· | ···················· | ···················· | ···················· |
| J | 6784 hCoV-19/England/MILK-99E46C/2020\|EPI_ISL_550592\|20 | ···················· | ···················· | ···················· | ···················· | ···················· | ···················· |
| J | 6785 hCoV-19/England/MILK-99E0D1/2020\|EPI_ISL_550594\|20 | ···················· | ···················· | ···················· | ···················· | ···················· | ···············A···· |
| J | 6786 hCoV-19/England/MILK-99CA5D/2020\|EPI_ISL_550595\|20 | ···················· | ···················· | ···················· | ···················· | ···················· | ···················· |
| J | 6787 hCoV-19/England/MILK-99DE14/2020\|EPI_ISL_550596\|20 | ···················· | ···················· | ···················· | ···················· | ···················· | ···················· |
| J | 6788 hCoV-19/England/MILK-99DCB0/2020\|EPI_ISL_550597\|20 | ···················· | ···················· | ···················· | ···················· | ···················· | ···················· |
| J | 6789 hCoV-19/England/MILK-99C7ED/2020\|EPI_ISL_550598\|20 | ···················· | ···················· | ···················· | ···················· | ···················· | ···················· |
| J | 6790 hCoV-19/England/CAMC-99134E/2020\|EPI_ISL_551851\|20 | ···················· | ·····C·············· | ···················· | ···················· | ···················· | ···················· |
| J | 6791 hCoV-19/England/CAMC-990FA5/2020\|EPI_ISL_551852\|20 | ···················· | ···················· | ···················· | ···················· | ···················· | ···················· |
| J | 6792 hCoV-19/England/CAMC-9911DB/2020\|EPI_ISL_551853\|20 | ···················· | ···················· | ···················· | ···················· | ···················· | ···················· |
| J | 6793 hCoV-19/England/CAMC-990989/2020\|EPI_ISL_551854\|20 | ···················· | ···················· | ···················· | ···················· | ···················· | ···················· |
| J | 6794 hCoV-19/England/CAMC-990DBA/2020\|EPI_ISL_551855\|20 | ···················· | ···················· | ···················· | ···················· | ···················· | ···················· |

| | | | | | | | |
|---|---|---|---|---|---|---|---|
| J | 6795 hCoV-19/England/CAMC-990A77/2020\|EPI_ISL_551856\|20 | ···················· | ···················· | ···················· | ···················· | ···················· | ···················· |
| J | 6796 hCoV-19/England/CAMC-9912E7/2020\|EPI_ISL_551857\|20 | ···················· | ···················· | ···················· | ···················· | ···················· | ···················· |
| J | 6797 hCoV-19/England/CAMC-990B29/2020\|EPI_ISL_551858\|20 | ···················· | ···················· | ···················· | ···················· | ···················· | ···················· |
| J | 6798 hCoV-19/England/CAMC-990A3B/2020\|EPI_ISL_551859\|20 | ···················· | ···················· | ···················· | ···················· | ···················· | ···················· |
| J | 6799 hCoV-19/England/CAMC-990A4A/2020\|EPI_ISL_551860\|20 | ···················· | ···················· | ···················· | ···················· | ···················· | ···················· |
| J | 6800 hCoV-19/England/CAMC-990C17/2020\|EPI_ISL_551861\|20 | ···················· | ···················· | ···················· | ···················· | ···················· | ···················· |
| J | 6801 hCoV-19/England/CAMC-990A2C/2020\|EPI_ISL_551862\|20 | ···················· | ···················· | ···················· | ···················· | ···················· | ···················· |
| J | 6802 hCoV-19/England/CAMC-991320/2020\|EPI_ISL_551863\|20 | ···················· | ···················· | ···················· | ···················· | ···················· | ···················· |
| J | 6803 hCoV-19/England/CAMC-9912AB/2020\|EPI_ISL_551864\|20 | ···················· | ···················· | ···················· | ···················· | ···················· | ···················· |
| J | 6804 hCoV-19/England/CAMC-990CDB/2020\|EPI_ISL_551865\|20 | ···················· | ···················· | ···················· | ···················· | ···················· | ···················· |
| J | 6805 hCoV-19/England/CAMC-990F87/2020\|EPI_ISL_551866\|20 | ···················· | ···················· | ···················· | ···················· | ···················· | ···················· |
| J | 6806 hCoV-19/England/CAMC-9908AA/2020\|EPI_ISL_551867\|20 | ···················· | ···················· | ···················· | ···················· | ···················· | ···················· |
| J | 6807 hCoV-19/England/CAMC-99092F/2020\|EPI_ISL_551868\|20 | ···················· | ···················· | ···················· | ···················· | ···················· | ···················· |
| J | 6808 hCoV-19/England/CAMC-990F4B/2020\|EPI_ISL_551869\|20 | ···················· | ···················· | ···················· | ···················· | ···················· | ···················· |
| J | 6809 hCoV-19/England/CAMC-9911AE/2020\|EPI_ISL_551870\|20 | ···················· | ···················· | ···················· | ···················· | ···················· | ···················· |
| J | 6810 hCoV-19/England/CAMC-9911BD/2020\|EPI_ISL_551871\|20 | ···················· | ···················· | ···················· | ···················· | ···················· | ···················· |
| J | 6811 hCoV-19/England/CAMC-9910A1/2020\|EPI_ISL_551872\|20 | ···················· | ···················· | ···················· | ···················· | ···················· | ···················· |
| J | 6812 hCoV-19/England/CAMC-990AB3/2020\|EPI_ISL_551873\|20 | ···················· | ·A·················· | ···················· | ···················· | ···················· | ···················· |
| J | 6813 hCoV-19/Northern Ireland/CAMC-990E6C/2020\|EPI_ISL_ | ···················· | ···················· | ···················· | ···················· | ···················· | ···················· |
| J | 6814 hCoV-19/England/CAMC-99086E/2020\|EPI_ISL_551876\|20 | ···················· | ···················· | ···················· | ···················· | ···················· | ···················· |
| J | 6815 hCoV-19/England/CAMC-99128D/2020\|EPI_ISL_551878\|20 | ···················· | ···················· | ···················· | ···················· | ···················· | ···················· |
| J | 6816 hCoV-19/England/CAMC-9910B0/2020\|EPI_ISL_551879\|20 | ···················· | ···················· | ···················· | ···················· | ···················· | ···················· |
| J | 6817 hCoV-19/England/CAMC-9909F2/2020\|EPI_ISL_551880\|20 | ···················· | ····A··············· | ···················· | ···················· | ···················· | ···················· |
| J | 6818 hCoV-19/England/CAMC-99126F/2020\|EPI_ISL_551881\|20 | ···················· | ···················· | ···················· | ···················· | ······T············· | ···················· |
| J | 6819 hCoV-19/England/CAMC-990CCC/2020\|EPI_ISL_551882\|20 | ···················· | ···················· | ···················· | ···················· | ······T············· | ···················· |
| J | 6820 hCoV-19/England/CAMC-990DF6/2020\|EPI_ISL_551883\|20 | ···················· | ···················· | ···················· | ···················· | ···················· | ···················· |
| J | 6821 hCoV-19/England/CAMC-9907DA/2020\|EPI_ISL_551884\|20 | ···················· | ···················· | ···················· | ···················· | ···················· | ···················· |
| J | 6822 hCoV-19/England/CAMC-990E02/2020\|EPI_ISL_551885\|20 | ···················· | ···················· | ···················· | ···················· | ···················· | ···················· |
| J | 6823 hCoV-19/England/CAMC-990F78/2020\|EPI_ISL_551886\|20 | ···················· | ···················· | ···················· | ···················· | ···················· | ···················· |
| J | 6824 hCoV-19/England/CAMC-99085F/2020\|EPI_ISL_551889\|20 | ···················· | ···················· | ···················· | ···················· | ···················· | ···················· |
| J | 6825 hCoV-19/England/CAMC-990C26/2020\|EPI_ISL_551890\|20 | ···················· | ·A·················· | ···················· | ···················· | ···················· | ···················· |
| J | 6826 hCoV-19/England/CAMC-991047/2020\|EPI_ISL_551891\|20 | ···················· | ···················· | ···················· | ···················· | ···················· | ···················· |
| J | 6827 hCoV-19/England/CAMC-9910DE/2020\|EPI_ISL_551892\|20 | ···················· | ····A··············· | ···················· | ···················· | ···················· | ···················· |
| J | 6828 hCoV-19/England/CAMC-990C08/2020\|EPI_ISL_551893\|20 | ···················· | ···················· | ···················· | ···················· | ···················· | ···················· |
| J | 6829 hCoV-19/England/CAMC-991108/2020\|EPI_ISL_551894\|20 | ···················· | ···················· | ···················· | ···················· | ···················· | ···············A···· |
| J | 6830 hCoV-19/England/CAMC-990831/2020\|EPI_ISL_551895\|20 | ···················· | ···················· | ···················· | ···················· | ···················· | ···················· |
| J | 6831 hCoV-19/England/CAMC-990D14/2020\|EPI_ISL_551898\|20 | ···················· | ···················· | ···················· | ···················· | ···················· | ···················· |
| J | 6832 hCoV-19/England/CAMC-990804/2020\|EPI_ISL_551899\|20 | ···················· | ···················· | ···················· | ···················· | ···················· | ···················· |
| J | 6833 hCoV-19/England/ALDP-952CF3/2020\|EPI_ISL_554039\|20 | ···················· | ···················· | ···················· | ···················· | ···················· | ···················· |
| J | 6834 hCoV-19/England/ALDP-955DB1/2020\|EPI_ISL_554040\|20 | ···················· | ···················· | ···················· | ···················· | ···················· | ···················· |
| J | 6835 hCoV-19/England/ALDP-952507/2020\|EPI_ISL_554041\|20 | ···················· | ···················· | ···················· | ···················· | ···················· | ···················· |
| J | 6836 hCoV-19/Wales/MILK-95772A/2020\|EPI_ISL_554042\|2020 | ···················· | ···················· | ···················· | ···················· | ···················· | ···················· |
| J | 6837 hCoV-19/England/ALDP-954769/2020\|EPI_ISL_555370\|20 | ···················· | ·A·················· | ···················· | ···················· | ···················· | ···················· |
| J | 6838 hCoV-19/Wales/ALDP-952C4E/2020\|EPI_ISL_555371\|2020 | ···················· | ···················· | ···················· | ···················· | ···················· | ···················· |
| J | 6839 hCoV-19/England/ALDP-94E638/2020\|EPI_ISL_555372\|20 | ···················· | ···················· | ···················· | ···················· | ···················· | ···················· |
| J | 6840 hCoV-19/Wales/ALDP-94DFB6/2020\|EPI_ISL_555373\|2020 | ···················· | ···················· | ···················· | ···················· | ···················· | ···················· |
| J | 6841 hCoV-19/England/ALDP-9519B1/2020\|EPI_ISL_555374\|20 | ···················· | ···················· | ···················· | ···················· | ···················· | ···················· |
| J | 6842 hCoV-19/England/ALDP-94E629/2020\|EPI_ISL_555401\|20 | ···················· | ···················· | ···················· | ···················· | ···················· | ···················· |
| J | 6843 hCoV-19/England/ALDP-94E0EF/2020\|EPI_ISL_555402\|20 | ···················· | ···················· | ···················· | ···················· | ···················· | ···················· |
| J | 6844 hCoV-19/England/ALDP-94E692/2020\|EPI_ISL_555403\|20 | ···················· | ···················· | ···················· | ···················· | ···················· | ···················· |
| J | 6845 hCoV-19/England/ALDP-95474B/2020\|EPI_ISL_555404\|20 | ···················· | ···················· | ···················· | ···················· | ···················· | ···················· |
| J | 6846 hCoV-19/England/ALDP-94E4D4/2020\|EPI_ISL_555405\|20 | ···················· | ···················· | ···················· | ···················· | ···················· | ···················· |
| J | 6847 hCoV-19/Wales/ALDP-952C5D/2020\|EPI_ISL_555406\|2020 | ···················· | ···················· | ···················· | ···················· | ···················· | ···················· |
| J | 6848 hCoV-19/England/ALDP-954945/2020\|EPI_ISL_555407\|20 | ···················· | ···················· | ···················· | ···················· | ···················· | ···················· |
| J | 6849 hCoV-19/England/ALDP-94E45C/2020\|EPI_ISL_555408\|20 | ···················· | ···················· | ···················· | ···················· | ···················· | ···················· |
| J | 6850 hCoV-19/England/ALDP-952C99/2020\|EPI_ISL_555409\|20 | ···················· | ···················· | ···················· | ···················· | ···················· | ···················· |
| J | 6851 hCoV-19/England/ALDP-94E4B6/2020\|EPI_ISL_555410\|20 | ···················· | ···················· | ···················· | ···················· | ···················· | ···················· |
| J | 6852 hCoV-19/England/ALDP-94E37D/2020\|EPI_ISL_555412\|20 | ···················· | ···················· | ···················· | ···················· | ···················· | ···················· |
| J | 6853 hCoV-19/Wales/ALDP-954936/2020\|EPI_ISL_555413\|2020 | ···················· | ···················· | ···················· | ···················· | ···················· | ···················· |
| J | 6854 hCoV-19/England/CAMC-94C324/2020\|EPI_ISL_556701\|20 | ···················· | ···················· | ···················· | ···················· | ···················· | ···················· |
| J | 6855 hCoV-19/England/CAMC-94AD0A/2020\|EPI_ISL_556702\|20 | ···················· | ···················· | ···················· | ···················· | ···················· | ···················· |
| J | 6856 hCoV-19/England/CAMC-94BF21/2020\|EPI_ISL_556704\|20 | ···················· | ···················· | ···················· | ···················· | ···················· | ···················· |
| J | 6857 hCoV-19/England/CAMC-94658E/2020\|EPI_ISL_556705\|20 | ···················· | ······A············· | ···················· | ···················· | ···················· | ···················· |
| J | 6858 hCoV-19/England/CAMC-94AA9A/2020\|EPI_ISL_556706\|20 | ···················· | ···················· | ···················· | ···················· | ···················· | ···················· |
| J | 6859 hCoV-19/England/CAMC-94C245/2020\|EPI_ISL_556707\|20 | ···················· | ···················· | ···················· | ···················· | ···················· | ···················· |
| J | 6860 hCoV-19/England/CAMC-945F54/2020\|EPI_ISL_556708\|20 | ···················· | ···················· | ···················· | ···················· | ···················· | ···················· |
| J | 6861 hCoV-19/Wales/CAMC-945BBA/2020\|EPI_ISL_556709\|2020 | ···················· | ···················· | ···················· | ···················· | ···················· | ···················· |
| J | 6862 hCoV-19/England/CAMC-94B237/2020\|EPI_ISL_556710\|20 | ···················· | ·A·················· | ···················· | ···················· | ···················· | ···A················ |
| J | 6863 hCoV-19/England/CAMC-94631B/2020\|EPI_ISL_556711\|20 | ···················· | ···················· | ···················· | ···················· | ···················· | ···················· |
| J | 6864 hCoV-19/England/CAMC-946542/2020\|EPI_ISL_556712\|20 | ···················· | ·A·················· | ···················· | ···················· | ···················· | ···················· |
| J | 6865 hCoV-19/England/CAMC-945E2A/2020\|EPI_ISL_556713\|20 | ···················· | ···················· | ···················· | ···················· | ···················· | ···················· |
| J | 6866 hCoV-19/England/CAMC-945B6F/2020\|EPI_ISL_556714\|20 | ···················· | ···················· | ···················· | ···················· | ···················· | ···················· |
| J | 6867 hCoV-19/England/CAMC-94C175/2020\|EPI_ISL_556715\|20 | ···················· | ···················· | ···················· | ···················· | ···················· | ···················· |
| J | 6868 hCoV-19/England/CAMC-94BE9D/2020\|EPI_ISL_556716\|20 | ···················· | ···················· | ···················· | ···················· | ···················· | ···················· |
| J | 6869 hCoV-19/England/CAMC-945C02/2020\|EPI_ISL_556717\|20 | ···················· | ···················· | ···················· | ···················· | ···················· | ···················· |
| J | 6870 hCoV-19/England/CAMC-949F32/2020\|EPI_ISL_556718\|20 | ···················· | ···················· | ···················· | ···················· | ···················· | ···················· |
| J | 6871 hCoV-19/England/CAMC-946348/2020\|EPI_ISL_556720\|20 | ···················· | ···················· | ···················· | ···················· | ···················· | ···················· |
| J | 6872 hCoV-19/England/CAMC-946366/2020\|EPI_ISL_556722\|20 | ···················· | ···················· | ···················· | ···················· | ···················· | ···················· |
| J | 6873 hCoV-19/England/CAMC-9461C6/2020\|EPI_ISL_556723\|20 | ···················· | ···················· | ···················· | ···················· | ···················· | ···················· |
| J | 6874 hCoV-19/England/CAMC-946409/2020\|EPI_ISL_556724\|20 | ···················· | ···················· | ···················· | ···················· | ···················· | ···················· |

| | | | | | | | |
|---|---|---|---|---|---|---|---|
| J | 6875 hCoV-19/England/CAMC-94BD18/2020\|EPI_ISL_556726\|20 | ···················· | ·A·················· | ···················· | ···················· | ···················· | ···················· |
| J | 6876 hCoV-19/England/CAMC-945EFD/2020\|EPI_ISL_556727\|20 | ···················· | ···················· | ···················· | ···················· | ···················· | ···················· |
| J | 6877 hCoV-19/England/CAMC-94C360/2020\|EPI_ISL_556728\|20 | ···················· | ···················· | ···················· | ···················· | ···················· | ···················· |
| J | 6878 hCoV-19/England/CAMC-945B05/2020\|EPI_ISL_556729\|20 | ···················· | ···················· | ···················· | ···················· | ···················· | ···················· |
| J | 6879 hCoV-19/England/CAMC-9496F1/2020\|EPI_ISL_556730\|20 | ···················· | ···················· | ···················· | ···················· | ···················· | ···················· |
| J | 6880 hCoV-19/England/CAMC-945B7E/2020\|EPI_ISL_556731\|20 | ···················· | ···················· | ···················· | ···················· | ···················· | ···················· |
| J | 6881 hCoV-19/England/CAMC-945C5D/2020\|EPI_ISL_556732\|20 | ···················· | ·A·················· | ···················· | ···················· | ···················· | ···················· |
| J | 6882 hCoV-19/England/CAMC-94858C/2020\|EPI_ISL_556733\|20 | ···················· | ·A·················· | ···················· | ···················· | ···················· | ···················· |
| J | 6883 hCoV-19/England/CAMC-94A739/2020\|EPI_ISL_556734\|20 | ···················· | ···················· | ···················· | ···················· | ···················· | ···················· |
| J | 6884 hCoV-19/England/CAMC-94863E/2020\|EPI_ISL_556735\|20 | ···················· | ···················· | ···················· | ···················· | ···················· | ···················· |
| J | 6885 hCoV-19/England/CAMC-94912D/2020\|EPI_ISL_556736\|20 | ···················· | ······A············· | ···················· | ···················· | ···················· | ···················· |
| J | 6886 hCoV-19/England/CAMC-948B8A/2020\|EPI_ISL_556737\|20 | ···················· | ···················· | ···················· | ···················· | ···················· | ···················· |
| J | 6887 hCoV-19/England/CAMC-949FD8/2020\|EPI_ISL_556738\|20 | ···················· | ·A·················· | ···················· | ···················· | ···················· | ···················· |
| J | 6888 hCoV-19/England/CAMC-948A8D/2020\|EPI_ISL_556739\|20 | ···················· | ···················· | ···················· | ···················· | ···················· | ···················· |
| J | 6889 hCoV-19/England/CAMC-946B22/2020\|EPI_ISL_556740\|20 | ···················· | ···················· | ···················· | ···················· | ···················· | ···················· |
| J | 6890 hCoV-19/England/CAMC-94B9D8/2020\|EPI_ISL_556741\|20 | ···················· | ······A············· | ···················· | ···················· | ···················· | ···················· |
| J | 6891 hCoV-19/England/CAMC-94590B/2020\|EPI_ISL_556742\|20 | ···················· | ···················· | ···················· | ···················· | ···················· | ···················· |
| J | 6892 hCoV-19/England/CAMC-9451B8/2020\|EPI_ISL_556743\|20 | ···················· | ···················· | ···················· | ···················· | ···················· | ···················· |
| J | 6893 hCoV-19/England/CAMC-94BC1B/2020\|EPI_ISL_556744\|20 | ···················· | ···················· | ···················· | ···················· | ···················· | ···················· |
| J | 6894 hCoV-19/England/MILK-56133C/2020\|EPI_ISL_558300\|20 | ···················· | ···················· | ···················· | ···················· | ···················· | ···················· |
| J | 6895 hCoV-19/England/MILK-559C3C/2020\|EPI_ISL_558301\|20 | ···················· | ···················· | ···················· | ···················· | ···················· | ···················· |
| J | 6896 hCoV-19/England/MILK-564AF8/2020\|EPI_ISL_558302\|20 | ···················· | ···················· | ···················· | ···················· | ···················· | ···················· |
| J | 6897 hCoV-19/England/MILK-576F7C/2020\|EPI_ISL_558303\|20 | ···················· | ···················· | ···················· | ···················· | ···················· | ···················· |
| J | 6898 hCoV-19/England/MILK-56381D/2020\|EPI_ISL_558304\|20 | ···················· | ···················· | ···················· | ···················· | ···················· | ···················· |
| J | 6899 hCoV-19/England/MILK-55ED52/2020\|EPI_ISL_558306\|20 | ···················· | ···················· | ···················· | ···················· | ···················· | ···················· |
| J | 6900 hCoV-19/England/ALDP-4A94FA/2020\|EPI_ISL_559617\|20 | ···················· | ···················· | ···················· | ···················· | ···················· | ···················· |
| J | 6901 hCoV-19/England/ALDP-4A392E/2020\|EPI_ISL_559618\|20 | ···················· | ···················· | ···················· | ···················· | ···················· | ···················· |
| J | 6902 hCoV-19/Wales/ALDP-4AC2C0/2020\|EPI_ISL_559619\|2020 | ···················· | ···················· | ···················· | ···················· | ···················· | ···················· |
| J | 6903 hCoV-19/England/ALDP-4A76C9/2020\|EPI_ISL_559627\|20 | ···················· | ···················· | ···················· | ···················· | ···················· | ···················· |
| J | 6904 hCoV-19/England/ALDP-4A8868/2020\|EPI_ISL_559629\|20 | ···················· | ···················· | ···················· | ···················· | ···················· | ···················· |
| J | 6905 hCoV-19/England/ALDP-49F5B8/2020\|EPI_ISL_559630\|20 | ···················· | ···················· | ···················· | ···················· | ···················· | ···················· |
| J | 6906 hCoV-19/England/ALDP-49F6A6/2020\|EPI_ISL_559631\|20 | ···················· | ···················· | ···················· | ···················· | ···················· | ···················· |
| J | 6907 hCoV-19/England/ALDP-4AD915/2020\|EPI_ISL_559632\|20 | ···················· | ···················· | ···················· | ···················· | ···················· | ···················· |
| J | 6908 hCoV-19/England/ALDP-4A5EE1/2020\|EPI_ISL_559633\|20 | ···················· | ···················· | ···················· | ···················· | ···················· | ···················· |
| J | 6909 hCoV-19/England/ALDP-4A4D6D/2020\|EPI_ISL_559634\|20 | ···················· | ···················· | ···················· | ···················· | ···················· | ···················· |
| J | 6910 hCoV-19/England/ALDP-4A822E/2020\|EPI_ISL_559635\|20 | ···················· | ···················· | ···················· | ···················· | ···················· | ···················· |
| J | 6911 hCoV-19/England/ALDP-4A5722/2020\|EPI_ISL_559636\|20 | ···················· | ···················· | ···················· | ···················· | ···················· | ···················· |
| J | 6912 hCoV-19/England/ALDP-4A77E4/2020\|EPI_ISL_559637\|20 | ···················· | ···················· | ···················· | ···················· | ···················· | ···················· |
| J | 6913 hCoV-19/Sweden/RV-FOI-9/2020\|EPI_ISL_548962\|2020-0 | ···················· | ···················· | ···················· | ···················· | ···················· | ···················· |
| J | 6914 hCoV-19/Sweden/RV-FOI-10/2020\|EPI_ISL_548963\|2020- | ···················· | ···················· | ···················· | ···················· | ···················· | ···················· |
| J | 6915 hCoV-19/United Arab Emirates/DU-A-S13-3_S8/2020\|EP | ···················· | ···················· | ···················· | ···················· | ···················· | ···················· |
| J | 6916 hCoV-19/United Arab Emirates/DU-A-S2-3_S2/2020\|EPI | ···················· | ···················· | ···················· | ···················· | ···················· | ···················· |
| J | 6917 hCoV-19/Singapore/971/2020\|EPI_ISL_548979\|2020-03- | ···················· | ·A·················· | ···················· | ···················· | ···················· | ···················· |
| J | 6918 hCoV-19/Singapore/978/2020\|EPI_ISL_548986\|2020-03- | ···················· | ···················· | ···················· | ···················· | ···················· | ···················· |
| J | 6919 hCoV-19/Singapore/979/2020\|EPI_ISL_548987\|2020-03- | ···················· | ···················· | ···················· | ···················· | ···················· | ···················· |
| J | 6920 hCoV-19/Singapore/980/2020\|EPI_ISL_548988\|2020-03- | ···················· | ···················· | ···················· | ···················· | ···················· | ···················· |
| J | 6921 hCoV-19/Singapore/981/2020\|EPI_ISL_548989\|2020-03- | ···················· | ···················· | ···················· | ···················· | ···················· | ···················· |
| J | 6922 hCoV-19/Singapore/982/2020\|EPI_ISL_548990\|2020-03- | ···················· | ···················· | ···················· | ···················· | ···················· | ···················· |
| J | 6923 hCoV-19/Singapore/983/2020\|EPI_ISL_548991\|2020-03- | ···················· | ···················· | ···················· | ···················· | ···················· | ···················· |
| J | 6924 hCoV-19/Singapore/984/2020\|EPI_ISL_548992\|2020-03- | ···················· | ···················· | ···················· | ···················· | ···················· | ···················· |
| J | 6925 hCoV-19/Singapore/985/2020\|EPI_ISL_548993\|2020-03- | ···················· | ···················· | ···················· | ···················· | ···················· | ···················· |
| J | 6926 hCoV-19/Singapore/986/2020\|EPI_ISL_548994\|2020-03- | ···················· | ···················· | ···················· | ···················· | ···················· | ···················· |
| J | 6927 hCoV-19/Singapore/987/2020\|EPI_ISL_548995\|2020-03- | ···················· | ···················· | ···················· | ···················· | ···················· | ···················· |
| J | 6928 hCoV-19/Singapore/988/2020\|EPI_ISL_548996\|2020-03- | ···················· | ···················· | ···················· | ···················· | ···················· | ···················· |
| J | 6929 hCoV-19/Singapore/989/2020\|EPI_ISL_548997\|2020-03- | ···················· | ···················· | ···················· | ···················· | ···················· | ···················· |
| J | 6930 hCoV-19/Singapore/990/2020\|EPI_ISL_548998\|2020-03- | ···················· | ···················· | ···················· | ···················· | ···················· | ···················· |
| J | 6931 hCoV-19/Singapore/991/2020\|EPI_ISL_548999\|2020-03- | ···················· | ···················· | ···················· | ···················· | ···················· | ···················· |
| J | 6932 hCoV-19/England/MILK-9931AC/2020\|EPI_ISL_552147\|20 | ···················· | ···················· | ···················· | ···················· | ···················· | ···················· |
| J | 6933 hCoV-19/England/MILK-9927E7/2020\|EPI_ISL_552149\|20 | ···················· | ···················· | ···················· | ···················· | ······T············· | ···················· |
| J | 6934 hCoV-19/England/MILK-9934FE/2020\|EPI_ISL_552150\|20 | ···················· | ···················· | ···················· | ···················· | ···················· | ···················· |
| J | 6935 hCoV-19/England/CAMC-991575/2020\|EPI_ISL_552151\|20 | ···················· | ···················· | ···················· | ···················· | ···················· | ···················· |
| J | 6936 hCoV-19/England/MILK-9932F4/2020\|EPI_ISL_552152\|20 | ···················· | ···················· | ···················· | ···················· | ···················· | ···················· |
| J | 6937 hCoV-19/England/MILK-9929B4/2020\|EPI_ISL_552153\|20 | ···················· | ···················· | ···················· | ···················· | ···················· | ···················· |
| J | 6938 hCoV-19/England/MILK-992CBB/2020\|EPI_ISL_552154\|20 | ···················· | ···················· | ···················· | ···················· | ···················· | ···················· |
| J | 6939 hCoV-19/England/MILK-992F3A/2020\|EPI_ISL_552155\|20 | ···················· | ···················· | ···················· | ···················· | ···················· | ···················· |
| J | 6940 hCoV-19/England/MILK-992A93/2020\|EPI_ISL_552156\|20 | ···················· | ···················· | ···················· | ···················· | ···················· | ···················· |
| J | 6941 hCoV-19/England/MILK-992B54/2020\|EPI_ISL_552158\|20 | ···················· | ···················· | ···················· | ···················· | ···················· | ···················· |
| J | 6942 hCoV-19/England/MILK-993018/2020\|EPI_ISL_552159\|20 | ···················· | ·A·················· | ···················· | ···················· | ···················· | ···················· |
| J | 6943 hCoV-19/England/MILK-993160/2020\|EPI_ISL_552161\|20 | ···················· | ···················· | ···················· | ···················· | ···················· | ···················· |
| J | 6944 hCoV-19/Wales/MILK-992AB1/2020\|EPI_ISL_552162\|2020 | ···················· | ·A·················· | ···················· | ···················· | ···················· | ···A················ |
| J | 6945 hCoV-19/England/MILK-992F1C/2020\|EPI_ISL_552163\|20 | ···················· | ···················· | ···················· | ···················· | ···················· | ···················· |
| J | 6946 hCoV-19/Wales/MILK-992820/2020\|EPI_ISL_552164\|2020 | ···················· | ···················· | ···················· | ···················· | ···················· | ···················· |
| J | 6947 hCoV-19/England/MILK-992C42/2020\|EPI_ISL_552165\|20 | ···················· | ···················· | ···················· | ···················· | ···················· | ···················· |
| J | 6948 hCoV-19/England/CAMC-991584/2020\|EPI_ISL_552166\|20 | ···················· | ···················· | ···················· | ···················· | ···················· | ···················· |
| J | 6949 hCoV-19/England/MILK-993036/2020\|EPI_ISL_552167\|20 | ···················· | ···················· | ···················· | ···················· | ···················· | ···················· |
| J | 6950 hCoV-19/England/MILK-992E2E/2020\|EPI_ISL_552168\|20 | ···················· | ·A·················· | ···················· | ···················· | ···················· | ···A················ |
| J | 6951 hCoV-19/England/MILK-992F0D/2020\|EPI_ISL_552169\|20 | ···················· | ···················· | ···················· | ···················· | ···················· | ···················· |
| J | 6952 hCoV-19/England/CAMC-99151B/2020\|EPI_ISL_552170\|20 | ···················· | ···················· | ···················· | ···················· | ···················· | ···················· |
| J | 6953 hCoV-19/England/MILK-992F76/2020\|EPI_ISL_552171\|20 | ···················· | ···················· | ···················· | ···················· | ···················· | ···················· |
| J | 6954 hCoV-19/England/MILK-9928B7/2020\|EPI_ISL_552172\|20 | ···················· | ·A·················· | ···················· | ···················· | ···················· | ···················· |

| | | | | | | | | |
|---|---|---|---|---|---|---|---|---|
| J | 6955 | hCoV-19/England/MILK-991E7A/2020\|EPI_ISL_552174\|20 | ···················· | ···················· | ···················· | ···················· | ···················· | ···················· |
| J | 6956 | hCoV-19/England/MILK-99335B/2020\|EPI_ISL_552175\|20 | ···················· | ···················· | ···················· | ···················· | ···················· | ···················· |
| J | 6957 | hCoV-19/England/MILK-9926CC/2020\|EPI_ISL_552176\|20 | ···················· | ···················· | ···················· | ···················· | ···················· | ···················· |
| J | 6958 | hCoV-19/England/MILK-992A75/2020\|EPI_ISL_552177\|20 | ···················· | ···················· | ···················· | ···················· | ···················· | ···················· |
| J | 6959 | hCoV-19/England/MILK-991E89/2020\|EPI_ISL_552178\|20 | ···················· | ···················· | ···················· | ···················· | ···················· | ···················· |
| J | 6960 | hCoV-19/England/MILK-99269F/2020\|EPI_ISL_552179\|20 | ···················· | ···················· | ···················· | ···················· | ···················· | ···················· |
| J | 6961 | hCoV-19/England/CAMC-9914E1/2020\|EPI_ISL_552180\|20 | ···················· | ···················· | ···················· | ···················· | ···················· | ···················· |
| J | 6962 | hCoV-19/England/MILK-99334C/2020\|EPI_ISL_552181\|20 | ···················· | ···················· | ···················· | ···················· | ···················· | ···················· |
| J | 6963 | hCoV-19/Wales/MILK-992BCD/2020\|EPI_ISL_552182\|2020 | ···················· | ···················· | ···················· | ···················· | ···················· | ···················· |
| J | 6964 | hCoV-19/England/MILK-9929F0/2020\|EPI_ISL_552183\|20 | ···················· | ·A·······A·········· | ···················· | ···················· | ···················· | ···················· |
| J | 6965 | hCoV-19/England/CAMC-991663/2020\|EPI_ISL_552185\|20 | ···················· | ···················· | ···················· | ···················· | ···················· | ···················· |
| J | 6966 | hCoV-19/England/MILK-9929C3/2020\|EPI_ISL_552186\|20 | ···················· | ···················· | ···················· | ···················· | ···················· | ···················· |
| J | 6967 | hCoV-19/England/MILK-99468E/2020\|EPI_ISL_552188\|20 | ···················· | ···················· | ···················· | ···················· | ···················· | ···················· |
| J | 6968 | hCoV-19/England/MILK-995429/2020\|EPI_ISL_552189\|20 | ···················· | ···················· | ···················· | ···················· | ···················· | ···················· |
| J | 6969 | hCoV-19/Wales/MILK-994C31/2020\|EPI_ISL_552191\|2020 | ···················· | ···················· | ···················· | ···················· | ···················· | ···················· |
| J | 6970 | hCoV-19/England/MILK-9938F2/2020\|EPI_ISL_552192\|20 | ···················· | ···················· | ···················· | ···················· | ···················· | ···················· |
| J | 6971 | hCoV-19/England/MILK-991D04/2020\|EPI_ISL_552193\|20 | ···················· | ···················· | ···················· | ···················· | ···················· | ···················· |
| J | 6972 | hCoV-19/England/MILK-991DC8/2020\|EPI_ISL_552194\|20 | ···················· | ···················· | ···················· | ···················· | ···················· | ···················· |
| J | 6973 | hCoV-19/England/MILK-9949B2/2020\|EPI_ISL_552195\|20 | ···················· | ···················· | ···················· | ···················· | ···················· | ···················· |
| J | 6974 | hCoV-19/England/MILK-995052/2020\|EPI_ISL_552196\|20 | ···················· | ···················· | ···················· | ···················· | ···················· | ···················· |
| J | 6975 | hCoV-19/England/MILK-995465/2020\|EPI_ISL_552197\|20 | ···················· | ···················· | ···················· | ···················· | ···················· | ···················· |
| J | 6976 | hCoV-19/England/MILK-994B34/2020\|EPI_ISL_552198\|20 | ···················· | ···················· | ···················· | ···················· | ···················· | ···················· |
| J | 6977 | hCoV-19/England/ALDP-95A824/2020\|EPI_ISL_553478\|20 | ···················· | ···················· | ···················· | ···················· | ···················· | ···················· |
| J | 6978 | hCoV-19/England/ALDP-9555B9/2020\|EPI_ISL_553479\|20 | ···················· | ···················· | ···················· | ···················· | ···················· | ···················· |
| J | 6979 | hCoV-19/England/ALDP-954D1C/2020\|EPI_ISL_553480\|20 | ···················· | ···················· | ···················· | ···················· | ···················· | ···················· |
| J | 6980 | hCoV-19/England/ALDP-95CBCF/2020\|EPI_ISL_553482\|20 | ···················· | ···················· | ···················· | ···················· | ···················· | ···················· |
| J | 6981 | hCoV-19/England/ALDP-954C00/2020\|EPI_ISL_553483\|20 | ···················· | ···················· | ···················· | ···················· | ···················· | ···················· |
| J | 6982 | hCoV-19/England/ALDP-95D0B0/2020\|EPI_ISL_553484\|20 | ···················· | ···················· | ···················· | ···················· | ···················· | ···················· |
| J | 6983 | hCoV-19/England/ALDP-95A03B/2020\|EPI_ISL_553486\|20 | ···················· | ···················· | ···················· | ···················· | ···················· | ···················· |
| J | 6984 | hCoV-19/England/ALDP-954AE8/2020\|EPI_ISL_553487\|20 | ···················· | ···················· | ···················· | ···················· | ···················· | ···················· |
| J | 6985 | hCoV-19/England/ALDP-95506D/2020\|EPI_ISL_553488\|20 | ···················· | ···················· | ···················· | ···················· | ···················· | ···················· |
| J | 6986 | hCoV-19/England/ALDP-955391/2020\|EPI_ISL_553489\|20 | ···················· | ···················· | ···················· | ···················· | ···················· | ···················· |
| J | 6987 | hCoV-19/England/ALDP-95A727/2020\|EPI_ISL_553491\|20 | ···················· | ···················· | ···················· | ···················· | ···················· | ···················· |
| J | 6988 | hCoV-19/England/ALDP-959F20/2020\|EPI_ISL_553492\|20 | ···················· | ···················· | ···················· | ···················· | ···················· | ···················· |
| J | 6989 | hCoV-19/England/ALDP-9549CD/2020\|EPI_ISL_553493\|20 | ···················· | ···················· | ···················· | ···················· | ···················· | ···················· |
| J | 6990 | hCoV-19/England/ALDP-95A666/2020\|EPI_ISL_553494\|20 | ···················· | ···················· | ···················· | ···················· | ···················· | ···················· |
| J | 6991 | hCoV-19/England/ALDP-9549EB/2020\|EPI_ISL_553495\|20 | ···················· | ···················· | ···················· | ···················· | ···················· | ···················· |
| J | 6992 | hCoV-19/England/ALDP-95A52D/2020\|EPI_ISL_553496\|20 | ···················· | ···················· | ···················· | ···················· | ···················· | ···················· |
| J | 6993 | hCoV-19/England/ALDP-954BC7/2020\|EPI_ISL_553498\|20 | ···················· | ···················· | ···················· | ···················· | ···················· | ···················· |
| J | 6994 | hCoV-19/England/ALDP-95A1FC/2020\|EPI_ISL_553499\|20 | ···················· | ···················· | ···················· | ···················· | ···················· | ···················· |
| J | 6995 | hCoV-19/England/ALDP-95A596/2020\|EPI_ISL_553511\|20 | ···················· | ···················· | ···················· | ···················· | ···················· | ···················· |
| J | 6996 | hCoV-19/Wales/ALDP-95C9C5/2020\|EPI_ISL_553512\|2020 | ···················· | ···················· | ···················· | ···················· | ···················· | ···················· |
| J | 6997 | hCoV-19/England/ALDP-95D180/2020\|EPI_ISL_553513\|20 | ···················· | ···················· | ···················· | ···················· | ···················· | ···················· |
| J | 6998 | hCoV-19/England/ALDP-95A3F6/2020\|EPI_ISL_553515\|20 | ···················· | ···················· | ···················· | ···················· | ···················· | ···················· |
| J | 6999 | hCoV-19/England/ALDP-95CF1E/2020\|EPI_ISL_553516\|20 | ···················· | ···················· | ···················· | ···················· | ···················· | ···················· |
| J | 7000 | hCoV-19/England/ALDP-9554DA/2020\|EPI_ISL_553517\|20 | ···················· | ···················· | ···················· | ···················· | ···················· | ···················· |
| J | 7001 | hCoV-19/England/ALDP-95A350/2020\|EPI_ISL_553518\|20 | ···················· | ···················· | ···················· | ···················· | ···················· | ···················· |
| J | 7002 | hCoV-19/England/ALDP-95530A/2020\|EPI_ISL_553519\|20 | ···················· | ·A·················· | ···················· | ···················· | ···················· | ···················· |
| J | 7003 | hCoV-19/England/ALDP-95504F/2020\|EPI_ISL_553521\|20 | ···················· | ···················· | ···················· | ···················· | ···················· | ···················· |
| J | 7004 | hCoV-19/England/MILK-95806A/2020\|EPI_ISL_554842\|20 | ···················· | ···················· | ···················· | ···················· | ···················· | ···················· |
| J | 7005 | hCoV-19/England/MILK-957F31/2020\|EPI_ISL_554843\|20 | ···················· | ···················· | ···················· | ···················· | ···················· | ···················· |
| J | 7006 | hCoV-19/England/MILK-95803D/2020\|EPI_ISL_554844\|20 | ···················· | ···················· | ···················· | ···················· | ···················· | ···················· |
| J | 7007 | hCoV-19/England/ALDP-94459F/2020\|EPI_ISL_554846\|20 | ···················· | ···················· | ···················· | ···················· | ···················· | ···················· |
| J | 7008 | hCoV-19/England/MILK-9584F5/2020\|EPI_ISL_554851\|20 | ···················· | ···················· | ···················· | ···················· | ···················· | ···················· |
| J | 7009 | hCoV-19/England/MILK-958A11/2020\|EPI_ISL_554852\|20 | ···················· | ···················· | ···················· | ···················· | ···················· | ···················· |
| J | 7010 | hCoV-19/England/ALDP-8E39F1/2020\|EPI_ISL_557001\|20 | ···················· | ···················· | ···················· | ···················· | ···················· | ···················· |
| J | 7011 | hCoV-19/England/ALDP-8D9BE9/2020\|EPI_ISL_557003\|20 | ···················· | ···················· | ···················· | ···················· | ···················· | ···················· |
| J | 7012 | hCoV-19/England/ALDP-8E153B/2020\|EPI_ISL_557004\|20 | ···················· | ···················· | ···················· | ···················· | ···················· | ···················· |
| J | 7013 | hCoV-19/England/ALDP-8DF6F1/2020\|EPI_ISL_557005\|20 | ···················· | ···················· | ···················· | ···················· | ···················· | ···················· |
| J | 7014 | hCoV-19/England/ALDP-8B465C/2020\|EPI_ISL_557006\|20 | ···················· | ···················· | ···················· | ···················· | ···················· | ···················· |
| J | 7015 | hCoV-19/England/ALDP-8BAD33/2020\|EPI_ISL_557007\|20 | ···················· | ···················· | ···················· | ···················· | ···················· | ···················· |
| J | 7016 | hCoV-19/England/ALDP-8D6205/2020\|EPI_ISL_557008\|20 | ···················· | ···················· | ···················· | ···················· | ···················· | ···················· |
| J | 7017 | hCoV-19/England/ALDP-8B9D16/2020\|EPI_ISL_557009\|20 | ···················· | ···················· | ···················· | ···················· | ···················· | ···················· |
| J | 7018 | hCoV-19/England/ALDP-8D4173/2020\|EPI_ISL_557010\|20 | ···················· | ···················· | ···················· | ···················· | ···················· | ···················· |
| J | 7019 | hCoV-19/England/ALDP-9149E7/2020\|EPI_ISL_557012\|20 | ···················· | ···················· | ···················· | ···················· | ···················· | ···················· |
| J | 7020 | hCoV-19/England/ALDP-8D71CB/2020\|EPI_ISL_557013\|20 | ···················· | ···················· | ···················· | ···················· | ···················· | ···················· |
| J | 7021 | hCoV-19/England/ALDP-8B42FE/2020\|EPI_ISL_557015\|20 | ···················· | ···················· | ···················· | ···················· | ···················· | ···················· |
| J | 7022 | hCoV-19/England/ALDP-8D9C8C/2020\|EPI_ISL_557017\|20 | ···················· | ···················· | ···················· | ···················· | ···················· | ···················· |
| J | 7023 | hCoV-19/England/ALDP-8B7F9A/2020\|EPI_ISL_557018\|20 | ···················· | ···················· | ···················· | ···················· | ···················· | ···················· |
| J | 7024 | hCoV-19/England/ALDP-8D3244/2020\|EPI_ISL_557019\|20 | ···················· | ···················· | ···················· | ···················· | ···················· | ···················· |
| J | 7025 | hCoV-19/England/ALDP-8D5549/2020\|EPI_ISL_557020\|20 | ···················· | ···················· | ···················· | ···················· | ···················· | ···················· |
| J | 7026 | hCoV-19/England/ALDP-8DDFBC/2020\|EPI_ISL_557022\|20 | ···················· | ···················· | ···················· | ···················· | ···················· | ···················· |
| J | 7027 | hCoV-19/England/ALDP-8D2290/2020\|EPI_ISL_557023\|20 | ···················· | ···················· | ···················· | ···················· | ···················· | ···················· |
| J | 7028 | hCoV-19/England/ALDP-8DE100/2020\|EPI_ISL_557024\|20 | ···················· | ···················· | ···················· | ···················· | ···················· | ···················· |
| J | 7029 | hCoV-19/England/ALDP-8B41B5/2020\|EPI_ISL_557025\|20 | ···················· | ···················· | ···················· | ···················· | ···················· | ···················· |
| J | 7030 | hCoV-19/England/ALDP-8E0E9B/2020\|EPI_ISL_557026\|20 | ···················· | ···················· | ···················· | ···················· | ···················· | ···················· |
| J | 7031 | hCoV-19/England/ALDP-8DE08B/2020\|EPI_ISL_557027\|20 | ···················· | ···················· | ···················· | ···················· | ···················· | ···················· |
| J | 7032 | hCoV-19/England/ALDP-8FF87D/2020\|EPI_ISL_557028\|20 | ···················· | ···················· | ···················· | ···················· | ···················· | ···················· |
| J | 7033 | hCoV-19/England/ALDP-8B9CFB/2020\|EPI_ISL_557029\|20 | ···················· | ···················· | ···················· | ···················· | ···················· | ···················· |
| J | 7034 | hCoV-19/England/ALDP-8FC7A0/2020\|EPI_ISL_557030\|20 | ···················· | ···················· | ···················· | ···················· | ···················· | ···················· |

| | | | | | | | |
|---|---|---|---|---|---|---|---|
| J | 7035 hCoV-19/England/ALDP-8D9DB6/2020\|EPI_ISL_557031\|20 | .................... | .................... | .................... | .................... | .................... | .................... |
| J | 7036 hCoV-19/England/ALDP-8D7592/2020\|EPI_ISL_557034\|20 | .................... | .................... | .................... | .................... | .................... | .................... |
| J | 7037 hCoV-19/England/ALDP-8D452C/2020\|EPI_ISL_557036\|20 | .................... | .................... | .................... | .................... | .................... | .................... |
| J | 7038 hCoV-19/England/ALDP-8B6A6D/2020\|EPI_ISL_557037\|20 | .................... | .................... | .................... | .................... | .................... | .................... |
| J | 7039 hCoV-19/England/ALDP-8B8922/2020\|EPI_ISL_557038\|20 | .................... | .................... | .................... | .................... | .................... | .................... |
| J | 7040 hCoV-19/England/ALDP-9015E9/2020\|EPI_ISL_557039\|20 | .................... | .................... | .................... | .................... | .................... | .................... |
| J | 7041 hCoV-19/England/ALDP-599DE2/2020\|EPI_ISL_558307\|20 | .................... | .................... | .................... | .................... | .................... | .................... |
| J | 7042 hCoV-19/England/MILK-5637C5/2020\|EPI_ISL_558309\|20 | .................... | .................... | .................... | .................... | .................... | .................... |
| J | 7043 hCoV-19/England/ALDP-59985A/2020\|EPI_ISL_558312\|20 | .................... | .................... | .................... | .................... | .................... | .................... |
| J | 7044 hCoV-19/England/MILK-5678CE/2020\|EPI_ISL_558313\|20 | .................... | .................... | .................... | .................... | .................... | .................... |
| J | 7045 hCoV-19/England/MILK-565AE8/2020\|EPI_ISL_558314\|20 | .................... | .................... | .................... | .................... | .................... | .................... |
| J | 7046 hCoV-19/England/MILK-562CA9/2020\|EPI_ISL_558315\|20 | .................... | .................... | .................... | .................... | .................... | .................... |
| J | 7047 hCoV-19/England/ALDP-599A09/2020\|EPI_ISL_558316\|20 | .................... | .................... | .................... | .................... | .................... | .................... |
| J | 7048 hCoV-19/England/MILK-576404/2020\|EPI_ISL_558317\|20 | .................... | .................... | .................... | .................... | .................... | .................... |
| J | 7049 hCoV-19/England/MILK-576ED9/2020\|EPI_ISL_558318\|20 | .................... | .................... | .................... | .................... | .................... | .................... |
| J | 7050 hCoV-19/England/MILK-57024C/2020\|EPI_ISL_558319\|20 | .................... | .................... | .................... | .................... | .................... | .................... |
| J | 7051 hCoV-19/England/MILK-55C2A0/2020\|EPI_ISL_558320\|20 | .................... | .................... | .................... | .................... | .................... | .................... |
| J | 7052 hCoV-19/England/ALDP-59B2E1/2020\|EPI_ISL_558321\|20 | .................... | .................... | .................... | .................... | .................... | .................... |
| J | 7053 hCoV-19/England/MILK-55949E/2020\|EPI_ISL_558322\|20 | .................... | .................... | .................... | .................... | .................... | .................... |
| J | 7054 hCoV-19/England/MILK-59625F/2020\|EPI_ISL_558323\|20 | .................... | .................... | .................... | .................... | .................... | .................... |
| J | 7055 hCoV-19/England/ALDP-599CA9/2020\|EPI_ISL_558324\|20 | .................... | .................... | .................... | .................... | .................... | .................... |
| J | 7056 hCoV-19/England/MILK-559DC0/2020\|EPI_ISL_558325\|20 | .................... | .................... | .................... | .................... | .................... | .................... |
| J | 7057 hCoV-19/England/MILK-56109F/2020\|EPI_ISL_558326\|20 | .................... | .................... | .................... | .................... | .................... | .................... |
| J | 7058 hCoV-19/England/MILK-566838/2020\|EPI_ISL_558327\|20 | .................... | .................... | .................... | .................... | .................... | .................... |
| J | 7059 hCoV-19/England/ALDP-59A7E3/2020\|EPI_ISL_558329\|20 | .................... | .................... | .................... | .................... | .................... | .................... |
| J | 7060 hCoV-19/England/MILK-5974EF/2020\|EPI_ISL_558331\|20 | .................... | .A.................. | .................... | .................... | .................... | .................... |
| J | 7061 hCoV-19/England/ALDP-59B0C9/2020\|EPI_ISL_558332\|20 | .................... | .................... | .................... | .................... | .................... | .................... |
| J | 7062 hCoV-19/Wales/MILK-56A8AD/2020\|EPI_ISL_558333\|2020 | .................... | .................... | .................... | .................... | .................... | .................... |
| J | 7063 hCoV-19/England/MILK-55E165/2020\|EPI_ISL_558334\|20 | .................... | .................... | .................... | .................... | .................... | .................... |
| J | 7064 hCoV-19/England/ALDP-59A992/2020\|EPI_ISL_558335\|20 | .................... | .................... | .................... | .................... | .................... | .................... |
| J | 7065 hCoV-19/England/MILK-55C079/2020\|EPI_ISL_558336\|20 | .................... | .................... | .................... | .................... | .................... | .................... |
| J | 7066 hCoV-19/England/MILK-565D2B/2020\|EPI_ISL_558337\|20 | .................... | .................... | .................... | .................... | .................... | .................... |
| J | 7067 hCoV-19/England/ALDP-5A01A0/2020\|EPI_ISL_558338\|20 | .................... | .................... | .................... | .................... | .................... | .................... |
| J | 7068 hCoV-19/England/MILK-55C1FE/2020\|EPI_ISL_558340\|20 | .................... | .................... | .................... | .................... | .................... | .................... |
| J | 7069 hCoV-19/England/MILK-563929/2020\|EPI_ISL_558341\|20 | .................... | .................... | .................... | .................... | .................... | .................... |
| J | 7070 hCoV-19/England/MILK-568D37/2020\|EPI_ISL_558342\|20 | .................... | .................... | .................... | .................... | .................... | .................... |
| J | 7071 hCoV-19/England/ALDP-599BD9/2020\|EPI_ISL_558343\|20 | .................... | .................... | .................... | .................... | .................... | .................... |
| J | 7072 hCoV-19/England/MILK-5753DB/2020\|EPI_ISL_558344\|20 | .................... | .................... | .................... | .................... | .................... | .................... |
| J | 7073 hCoV-19/England/ALDP-59A8C2/2020\|EPI_ISL_558345\|20 | .................... | .................... | .................... | .................... | .................... | .................... |
| J | 7074 hCoV-19/England/MILK-567A31/2020\|EPI_ISL_558347\|20 | .................... | .................... | .................... | .................... | .................... | .................... |
| J | 7075 hCoV-19/England/ALDP-59B4CD/2020\|EPI_ISL_558348\|20 | .................... | .................... | .................... | .................... | .................... | .................... |
| J | 7076 hCoV-19/England/ALDP-59A974/2020\|EPI_ISL_558350\|20 | .................... | .................... | .................... | .................... | .................... | .................... |
| J | 7077 hCoV-19/England/MILK-570446/2020\|EPI_ISL_558351\|20 | .................... | .................... | .................... | .................... | .................... | .................... |
| J | 7078 hCoV-19/England/MILK-58FB7C/2020\|EPI_ISL_558352\|20 | .................... | .................... | .................... | .................... | .................... | .................... |
| J | 7079 hCoV-19/England/MILK-56C4E3/2020\|EPI_ISL_558353\|20 | .................... | .................... | .................... | .................... | .................... | .................... |
| J | 7080 hCoV-19/England/ALDP-59AD69/2020\|EPI_ISL_558355\|20 | .................... | .................... | .................... | .................... | .................... | .................... |
| J | 7081 hCoV-19/England/MILK-599474/2020\|EPI_ISL_558357\|20 | .................... | .................... | .................... | .................... | .................... | .................... |
| J | 7082 hCoV-19/England/MILK-559CF0/2020\|EPI_ISL_558358\|20 | .................... | .................... | .................... | .................... | .................... | .................... |
| J | 7083 hCoV-19/Wales/MILK-598123/2020\|EPI_ISL_558359\|2020 | .................... | .................... | .................... | .................... | .................... | .................... |
| J | 7084 hCoV-19/England/MILK-56710F/2020\|EPI_ISL_558360\|20 | .................... | .................... | .................... | .................... | .................... | .................... |
| J | 7085 hCoV-19/England/MILK-566C5A/2020\|EPI_ISL_558361\|20 | .................... | .A.................. | .................... | .................... | .................... | ...A................ |
| J | 7086 hCoV-19/England/MILK-590BF0/2020\|EPI_ISL_558363\|20 | .................... | .................... | .................... | .................... | .................... | .................... |
| J | 7087 hCoV-19/England/ALDP-59C81B/2020\|EPI_ISL_558364\|20 | .................... | .................... | .................... | .................... | .................... | .................... |
| J | 7088 hCoV-19/England/MILK-56DF0F/2020\|EPI_ISL_558365\|20 | .................... | .................... | .................... | .................... | .................... | .................... |
| J | 7089 hCoV-19/England/MILK-578BC1/2020\|EPI_ISL_558367\|20 | .................... | .................... | .................... | .................... | .................... | .................... |
| J | 7090 hCoV-19/England/MILK-567D0B/2020\|EPI_ISL_558369\|20 | .................... | .................... | .................... | .................... | .................... | .................... |
| J | 7091 hCoV-19/England/ALDP-49E391/2020\|EPI_ISL_559600\|20 | .................... | .................... | .G.................. | .................... | .................... | .................... |
| J | 7092 hCoV-19/England/ALDP-49CD59/2020\|EPI_ISL_559601\|20 | .................... | .................... | .................... | .................... | .................... | .................... |
| J | 7093 hCoV-19/England/ALDP-4AB7FF/2020\|EPI_ISL_559602\|20 | .................... | .................... | .................... | .................... | .................... | .................... |
| J | 7094 hCoV-19/England/ALDP-4AD1B3/2020\|EPI_ISL_559603\|20 | .................... | .................... | .................... | .................... | .................... | .................... |
| J | 7095 hCoV-19/England/ALDP-49DBA9/2020\|EPI_ISL_559605\|20 | .................... | .................... | .................... | .................... | .................... | .................... |
| J | 7096 hCoV-19/England/ALDP-4A635A/2020\|EPI_ISL_559606\|20 | .................... | .................... | .................... | .................... | .................... | .................... |
| J | 7097 hCoV-19/England/ALDP-4A667F/2020\|EPI_ISL_559607\|20 | .................... | .................... | .................... | .................... | .................... | .................... |
| J | 7098 hCoV-19/England/ALDP-4A78C3/2020\|EPI_ISL_559608\|20 | .................... | .................... | .................... | .................... | .................... | .................... |
| J | 7099 hCoV-19/England/ALDP-49EF24/2020\|EPI_ISL_559609\|20 | .................... | .................... | .................... | .................... | .................... | .................... |
| J | 7100 hCoV-19/England/ALDP-49F80A/2020\|EPI_ISL_559610\|20 | .................... | .................... | .................... | .................... | .................... | .................... |
| J | 7101 hCoV-19/England/ALDP-49F3BE/2020\|EPI_ISL_559611\|20 | .................... | .................... | .................... | .................... | .................... | .................... |
| J | 7102 hCoV-19/England/ALDP-4A9B7D/2020\|EPI_ISL_559612\|20 | .................... | .................... | .................... | .................... | .................... | .................... |
| J | 7103 hCoV-19/England/ALDP-4A631E/2020\|EPI_ISL_559613\|20 | .................... | .................... | .................... | .................... | .................... | .................... |
| J | 7104 hCoV-19/England/ALDP-4A55CE/2020\|EPI_ISL_559614\|20 | .................... | .................... | .................... | .................... | .................... | .................... |
| J | 7105 hCoV-19/England/ALDP-4A9454/2020\|EPI_ISL_559615\|20 | .................... | .................... | .................... | .................... | .................... | .................... |
| J | 7106 hCoV-19/England/ALDP-4A91C6/2020\|EPI_ISL_559620\|20 | .................... | .................... | .................... | .................... | .................... | .................... |
| J | 7107 hCoV-19/England/ALDP-4A5889/2020\|EPI_ISL_559621\|20 | .................... | .................... | .................... | .................... | .................... | .................... |
| J | 7108 hCoV-19/England/ALDP-4ADF31/2020\|EPI_ISL_559622\|20 | .................... | .................... | .................... | .................... | .................... | .................... |
| J | 7109 hCoV-19/England/ALDP-497E79/2020\|EPI_ISL_559624\|20 | .................... | .................... | .................... | .................... | .................... | .................... |
| J | 7110 hCoV-19/England/ALDP-4A5634/2020\|EPI_ISL_559625\|20 | .................... | .................... | .................... | .................... | .................... | .................... |
| J | 7111 hCoV-19/England/ALDP-4A5B71/2020\|EPI_ISL_559626\|20 | .................... | .................... | .................... | .................... | .................... | .................... |
| J | 7112 hCoV-19/England/ALDP-49CE1A/2020\|EPI_ISL_559638\|20 | .................... | .................... | .................... | .................... | .................... | .................... |
| J | 7113 hCoV-19/England/ALDP-4A3D7D/2020\|EPI_ISL_559639\|20 | .................... | .................... | .................... | .................... | .................... | .................... |
| J | 7114 hCoV-19/England/ALDP-4A6E86/2020\|EPI_ISL_559640\|20 | .................... | .................... | .................... | .................... | .................... | .................... |

| | | | | | | | | | |
|---|---|---|---|---|---|---|---|---|---|
| J | 7115 | hCoV-19/England/ALDP-4A7492/2020\|EPI_ISL_559641\|20 | ···················· | ···················· | ···················· | ···················· | ···················· | ···················· | |
| J | 7116 | hCoV-19/England/ALDP-4A39F1/2020\|EPI_ISL_559642\|20 | ···················· | ···················· | ···················· | ···················· | ···················· | ···················· | |
| J | 7117 | hCoV-19/England/ALDP-4ADB00/2020\|EPI_ISL_559643\|20 | ···················· | ···················· | ···················· | ···················· | ···················· | ···················· | |
| J | 7118 | hCoV-19/England/ALDP-4A7D88/2020\|EPI_ISL_559645\|20 | ···················· | ···················· | ···················· | ···················· | ···················· | ···················· | |
| J | 7119 | hCoV-19/England/ALDP-4A717D/2020\|EPI_ISL_559646\|20 | ···················· | ···················· | ···················· | ···················· | ···················· | ···················· | |
| J | 7120 | hCoV-19/England/ALDP-497D12/2020\|EPI_ISL_559647\|20 | ···················· | ···················· | ···················· | ···················· | ···················· | ···················· | |
| J | 7121 | hCoV-19/England/ALDP-4ADDCE/2020\|EPI_ISL_559648\|20 | ···················· | ···················· | ···················· | ···················· | ···················· | ···················· | |
| J | 7122 | hCoV-19/England/ALDP-4AC363/2020\|EPI_ISL_559650\|20 | ···················· | ···················· | ···················· | ···················· | ···················· | ···················· | |
| J | 7123 | hCoV-19/England/ALDP-49EE63/2020\|EPI_ISL_559651\|20 | ···················· | ···················· | ···················· | ···················· | ···················· | ···················· | |
| J | 7124 | hCoV-19/England/ALDP-4A9A9E/2020\|EPI_ISL_559652\|20 | ···················· | ···················· | ···················· | ···················· | ···················· | ···················· | |
| J | 7125 | hCoV-19/England/ALDP-49F239/2020\|EPI_ISL_559653\|20 | ···················· | ···················· | ···················· | ···················· | ···················· | ···················· | |
| J | 7126 | hCoV-19/England/ALDP-4AD793/2020\|EPI_ISL_559654\|20 | ···················· | ···················· | ···················· | ···················· | ···················· | ···················· | |
| J | 7127 | hCoV-19/England/ALDP-4A93C0/2020\|EPI_ISL_559655\|20 | ···················· | ···················· | ···················· | ···················· | ···················· | ···················· | |
| J | 7128 | hCoV-19/England/ALDP-4A7EEF/2020\|EPI_ISL_559656\|20 | ···················· | ···················· | ···················· | ···················· | ···················· | ···················· | |
| J | 7129 | hCoV-19/England/ALDP-49C991/2020\|EPI_ISL_559657\|20 | ···················· | ···················· | ···················· | ···················· | ···················· | ···················· | |
| J | 7130 | hCoV-19/England/ALDP-4A6141/2020\|EPI_ISL_559658\|20 | ···················· | ···················· | ···················· | ···················· | ···················· | ···················· | |
| J | 7131 | hCoV-19/England/ALDP-4ABF24/2020\|EPI_ISL_559659\|20 | ···················· | ···················· | ···················· | ···················· | ···················· | ···················· | |
| J | 7132 | hCoV-19/England/ALDP-4A76BA/2020\|EPI_ISL_559660\|20 | ···················· | ···················· | ···················· | ···················· | ···················· | ···················· | |
| J | 7133 | hCoV-19/England/ALDP-4A869B/2020\|EPI_ISL_559661\|20 | ···················· | ···················· | ···················· | ···················· | ···················· | ···················· | |
| J | 7134 | hCoV-19/England/MILK-488CBD/2020\|EPI_ISL_559662\|20 | ···················· | ···················· | ···················· | ···················· | ···················· | ···················· | |
| J | 7135 | hCoV-19/England/MILK-482293/2020\|EPI_ISL_559663\|20 | ···················· | ···················· | ···················· | ···················· | ···················· | ···················· | |
| J | 7136 | hCoV-19/England/MILK-4887DA/2020\|EPI_ISL_559664\|20 | ···················· | ···················· | ···················· | ···················· | ···················· | ···················· | |
| J | 7137 | hCoV-19/England/MILK-487F79/2020\|EPI_ISL_559665\|20 | ···················· | ···················· | ···················· | ···················· | ···················· | ···················· | |
| J | 7138 | hCoV-19/England/MILK-488A4A/2020\|EPI_ISL_559666\|20 | ···················· | ···················A | ···················· | ···················· | ···················· | ···················· | 1 |
| J | 7139 | hCoV-19/England/MILK-483344/2020\|EPI_ISL_559667\|20 | ···················· | ···················· | ···················· | ···················· | ···················· | ···················· | |
| J | 7140 | hCoV-19/England/MILK-489214/2020\|EPI_ISL_559668\|20 | ···················· | ···················· | ···················· | ···················· | ···················· | ···················· | |
| J | 7141 | hCoV-19/England/MILK-48213C/2020\|EPI_ISL_559669\|20 | ···················· | ···················· | ···················· | ···················· | ···················· | ···················· | |
| J | 7142 | hCoV-19/England/MILK-48ADE5/2020\|EPI_ISL_559671\|20 | ···················· | ···················· | ···················· | ···················· | ···················· | ···················· | |
| J | 7143 | hCoV-19/England/MILK-483292/2020\|EPI_ISL_559672\|20 | ···················· | ···················· | ···················· | ···················· | ···················· | ···················· | |
| J | 7144 | hCoV-19/England/MILK-4822C0/2020\|EPI_ISL_559674\|20 | ···················· | ···················· | ···················· | ···················· | ···················· | ···················· | |
| J | 7145 | hCoV-19/England/MILK-4830B6/2020\|EPI_ISL_559675\|20 | ···················· | ···················· | ···················· | ···················· | ···················· | ···················· | |
| J | 7146 | hCoV-19/England/MILK-483238/2020\|EPI_ISL_559676\|20 | ···················· | ···················· | ···················· | ···················· | ···················· | ···················· | |
| J | 7147 | hCoV-19/England/MILK-486CA0/2020\|EPI_ISL_559677\|20 | ···················· | ···················· | ···················· | ···················· | ···················· | ···················· | |
| J | 7148 | hCoV-19/England/MILK-48548B/2020\|EPI_ISL_559678\|20 | ···················· | ···················· | ···················· | ···················· | ···················· | ···················· | |
| J | 7149 | hCoV-19/England/MILK-483186/2020\|EPI_ISL_559679\|20 | ···················· | ···················· | ···················· | ···················· | ···················· | ···················· | |
| J | 7150 | hCoV-19/England/MILK-4860D1/2020\|EPI_ISL_559680\|20 | ···················· | ···················· | ···················· | ···················· | ···················· | ···················· | |
| J | 7151 | hCoV-19/England/MILK-484826/2020\|EPI_ISL_559681\|20 | ···················· | ···················· | ···················· | ···················· | ···················· | ···················· | |
| J | 7152 | hCoV-19/England/MILK-48978E/2020\|EPI_ISL_559682\|20 | ···················· | ···················· | ···················· | ···················· | ···················· | ···················· | |
| J | 7153 | hCoV-19/England/MILK-483283/2020\|EPI_ISL_559683\|20 | ···················· | ···················· | ···················· | ···················· | ···················· | ···················· | |
| J | 7154 | hCoV-19/England/MILK-4882D9/2020\|EPI_ISL_559684\|20 | ···················· | ···················· | ···················· | ···················· | ···················· | ···················· | |
| J | 7155 | hCoV-19/England/MILK-48A34D/2020\|EPI_ISL_559685\|20 | ···················· | ···················· | ···················· | ···················· | ···················· | ···················· | |
| J | 7156 | hCoV-19/England/MILK-487A69/2020\|EPI_ISL_559686\|20 | ···················· | ···················· | ···················· | ···················· | ···················· | ···················· | |
| J | 7157 | hCoV-19/England/MILK-48249D/2020\|EPI_ISL_559687\|20 | ···················· | ···················· | ···················· | ···················· | ···················· | ···················· | |
| J | 7158 | hCoV-19/England/MILK-4882BB/2020\|EPI_ISL_559688\|20 | ···················· | ···················· | ···················· | ···················· | ···················· | ···················· | |
| J | 7159 | hCoV-19/England/MILK-4863C9/2020\|EPI_ISL_559689\|20 | ···················· | ···················· | ···················· | ···················· | ···················· | ···················· | |
| J | 7160 | hCoV-19/England/MILK-481FAC/2020\|EPI_ISL_559690\|20 | ···················· | ···················A | ···················· | ···················· | ···················· | ···················· | 1 |
| J | 7161 | hCoV-19/England/MILK-4862AE/2020\|EPI_ISL_559691\|20 | ···················· | ···················· | ···················· | ···················· | ···················· | ···················· | |
| J | 7162 | hCoV-19/England/MILK-485245/2020\|EPI_ISL_559692\|20 | ···················· | ···················· | ···················· | ···················· | ···················· | ···················· | |
| J | 7163 | hCoV-19/England/MILK-481443/2020\|EPI_ISL_559694\|20 | ···················· | ···················· | ···················· | ···················· | ···················· | ···················· | |
| J | 7164 | hCoV-19/England/MILK-481A50/2020\|EPI_ISL_559695\|20 | ···················· | ···················· | ···················· | ···················· | ···················· | ···················· | |
| J | 7165 | hCoV-19/England/MILK-482D1A/2020\|EPI_ISL_559696\|20 | ···················· | ···················· | ···················· | ···················· | ···················· | ···················· | |
| J | 7166 | hCoV-19/England/MILK-481829/2020\|EPI_ISL_559697\|20 | ···················· | ···················· | ···················· | ···················· | ···················· | ···················· | |
| J | 7167 | hCoV-19/England/MILK-485E14/2020\|EPI_ISL_559698\|20 | ···················· | ···················· | ···················· | ···················· | ···················· | ···················· | |
| J | 7168 | hCoV-19/England/MILK-4867CD/2020\|EPI_ISL_559699\|20 | ···················· | ···················· | ···················· | ···················· | ···················· | ···················· | |
| J | 7169 | hCoV-19/England/MILK-9954B0/2020\|EPI_ISL_552200\|20 | ···················· | ···················· | ···················· | ···················· | ···················· | ···················· | |
| J | 7170 | hCoV-19/England/MILK-99390E/2020\|EPI_ISL_552201\|20 | ···················· | ···················· | ···················· | ···················· | ······T············· | ···················· | |
| J | 7171 | hCoV-19/England/MILK-99519B/2020\|EPI_ISL_552203\|20 | ···················· | ·A·················· | ···················· | ···················· | ···················· | ···A················ | |
| J | 7172 | hCoV-19/England/MILK-995113/2020\|EPI_ISL_552204\|20 | ···················· | ···················· | ···················· | ···················· | ···················· | ···················· | |
| J | 7173 | hCoV-19/Wales/MILK-99476D/2020\|EPI_ISL_552205\|2020 | ···················· | ···················· | ···················· | ···················· | ···················· | ···················· | |
| J | 7174 | hCoV-19/England/MILK-9944EE/2020\|EPI_ISL_552206\|20 | ···················· | ···················· | ···················· | ···················· | ···················· | ···················· | |
| J | 7175 | hCoV-19/England/MILK-99531D/2020\|EPI_ISL_552208\|20 | ···················· | ···················· | ···················· | ···················· | ···················· | ···················· | |
| J | 7176 | hCoV-19/England/MILK-995104/2020\|EPI_ISL_552210\|20 | ···················· | ···················· | ···················· | ···················· | ···················· | ···················· | |
| J | 7177 | hCoV-19/England/MILK-994C7D/2020\|EPI_ISL_552211\|20 | ···················· | ···················· | ···················· | ···················· | ···················· | ···················· | |
| J | 7178 | hCoV-19/Wales/MILK-99475E/2020\|EPI_ISL_552212\|2020 | ···················· | ···················· | ···················· | ···················· | ···················· | ···················· | |
| J | 7179 | hCoV-19/England/MILK-994A82/2020\|EPI_ISL_552213\|20 | ···················· | ···················· | ···················· | ···················· | ···················· | ···················· | |
| J | 7180 | hCoV-19/England/MILK-994651/2020\|EPI_ISL_552214\|20 | ···················· | ···················· | ···················· | ···················· | ···················· | ···················· | |
| J | 7181 | hCoV-19/England/MILK-99533B/2020\|EPI_ISL_552215\|20 | ···················· | ···················· | ···················· | ···················· | ···················· | ···················· | |
| J | 7182 | hCoV-19/England/MILK-994EFF/2020\|EPI_ISL_552217\|20 | ···················· | ···················· | ···················· | ···················· | ···················· | ···················· | |
| J | 7183 | hCoV-19/England/MILK-9952C5/2020\|EPI_ISL_552218\|20 | ···················· | ·A·················· | ···················· | ···················· | ···················· | ···A················ | |
| J | 7184 | hCoV-19/England/MILK-993889/2020\|EPI_ISL_552219\|20 | ········T··········· | ···················· | ···················· | ···················· | ···················· | ···················· | |
| J | 7185 | hCoV-19/England/MILK-9948A6/2020\|EPI_ISL_552220\|20 | ···················· | ···················· | ···················· | ···················· | ···················· | ···················· | |
| J | 7186 | hCoV-19/England/MILK-9944B1/2020\|EPI_ISL_552221\|20 | ···················· | ···················· | ···················· | ···················· | ···················· | ···················· | |
| J | 7187 | hCoV-19/England/MILK-993C32/2020\|EPI_ISL_552222\|20 | ···················· | ···················· | ···················· | ···················· | ···················· | ···················· | |
| J | 7188 | hCoV-19/England/MILK-99526B/2020\|EPI_ISL_552223\|20 | ···················· | ···················· | ···················· | ···················· | ···················· | ···················· | |
| J | 7189 | hCoV-19/England/MILK-994AA0/2020\|EPI_ISL_552224\|20 | ···················· | ···················· | ···················· | ···················· | ···················· | ···················· | |
| J | 7190 | hCoV-19/England/MILK-9948B5/2020\|EPI_ISL_552225\|20 | ···················· | ······C············· | ···················· | ···················· | ···················· | ···················· | |
| J | 7191 | hCoV-19/England/MILK-993B26/2020\|EPI_ISL_552226\|20 | ···················· | ···················· | ···················· | ···················· | ···················· | ···················· | |
| J | 7192 | hCoV-19/England/MILK-991CF8/2020\|EPI_ISL_552228\|20 | ···················· | ···················· | ···················· | ···················· | ···················· | ···················· | |
| J | 7193 | hCoV-19/England/MILK-993B71/2020\|EPI_ISL_552229\|20 | ···················· | ···················· | ···················· | ···················· | ···················· | ···················· | |
| J | 7194 | hCoV-19/England/MILK-99534A/2020\|EPI_ISL_552230\|20 | ···················· | ···················· | ···················· | ···················· | ···················· | ···················· | |

| | | | | | | | |
|---|---|---|---|---|---|---|---|
| J | 7195 hCoV-19/England/MILK-994E95/2020\|EPI_ISL_552231\|20 | ···················· | ···················· | ····················· | ···················· | ···················· | ···················· |
| J | 7196 hCoV-19/England/MILK-9946AC/2020\|EPI_ISL_552232\|20 | ···················· | ···················· | ····················· | ···················· | ···················· | ···················· |
| J | 7197 hCoV-19/England/MILK-993C14/2020\|EPI_ISL_552234\|20 | ···················· | ···················· | ····················· | ···················· | ···················· | ···················· |
| J | 7198 hCoV-19/England/MILK-995386/2020\|EPI_ISL_552235\|20 | ···················· | ···················· | ····················· | ···················· | ···················· | ···················· |
| J | 7199 hCoV-19/England/MILK-991D8C/2020\|EPI_ISL_552236\|20 | ···················· | ···················· | ····················· | ···················· | ···················· | ···················· |
| J | 7200 hCoV-19/England/MILK-99517D/2020\|EPI_ISL_552237\|20 | ···················· | ···················· | ····················· | ···················· | ···················· | ···················· |
| J | 7201 hCoV-19/England/MILK-994536/2020\|EPI_ISL_552238\|20 | ···················· | ···················· | ····················· | ···················· | ···················· | ···················· |
| J | 7202 hCoV-19/England/MILK-994527/2020\|EPI_ISL_552241\|20 | ···················· | ···················· | ····················· | ···················· | ···················· | ···················· |
| J | 7203 hCoV-19/England/MILK-994F38/2020\|EPI_ISL_552242\|20 | ···················· | ···················· | ····················· | ···················· | ···················· | ···················· |
| J | 7204 hCoV-19/England/MILK-994BF8/2020\|EPI_ISL_552243\|20 | ···················· | ···················· | ····················· | ···················· | ·····T·············· | ···················· |
| J | 7205 hCoV-19/England/MILK-993B9F/2020\|EPI_ISL_552244\|20 | ···················· | ···················· | ····················· | ···················· | ···················· | ···················· |
| J | 7206 hCoV-19/England/MILK-9938D4/2020\|EPI_ISL_552245\|20 | ···················· | ···················· | ····················· | ···················· | ···················· | ···················· |
| J | 7207 hCoV-19/England/MILK-994C5F/2020\|EPI_ISL_552247\|20 | ···················· | ···················· | ····················· | ···················· | ···················· | ···················· |
| J | 7208 hCoV-19/England/MILK-995526/2020\|EPI_ISL_552248\|20 | ···················· | ···················· | ····················· | ···················· | ···················· | ···················· |
| J | 7209 hCoV-19/England/MILK-99483D/2020\|EPI_ISL_552249\|20 | ···················· | ·A·················· | ····················· | ···················· | ···················· | ··A················· |
| J | 7210 hCoV-19/England/MILK-9950F8/2020\|EPI_ISL_552251\|20 | ···················· | ···················· | ····················· | ···················· | ···················· | ···················· |
| J | 7211 hCoV-19/England/MILK-994DB6/2020\|EPI_ISL_552252\|20 | ···················· | ·A·················· | ····················· | ···················· | ···················· | ···················· |
| J | 7212 hCoV-19/England/MILK-994554/2020\|EPI_ISL_552253\|20 | ···················· | ·A·················· | ····················· | ···················· | ···················· | ··A················· |
| J | 7213 hCoV-19/England/MILK-995395/2020\|EPI_ISL_552254\|20 | ···················· | ···················· | ····················· | ···················· | ···················· | ···················· |
| J | 7214 hCoV-19/England/MILK-991CBC/2020\|EPI_ISL_552255\|20 | ···················· | ···················· | ····················· | ···················· | ···················· | ···················· |
| J | 7215 hCoV-19/England/MILK-99485B/2020\|EPI_ISL_552256\|20 | ···················· | ·A·················· | ····················· | ···················· | ···················· | ··A················· |
| J | 7216 hCoV-19/England/MILK-993898/2020\|EPI_ISL_552257\|20 | ···················· | ···················· | ····················· | ···················· | ···················· | ···················· |
| J | 7217 hCoV-19/England/MILK-9953C2/2020\|EPI_ISL_552260\|20 | ···················· | ···················· | ····················· | ···················· | ···················· | ···················· |
| J | 7218 hCoV-19/England/MILK-994484/2020\|EPI_ISL_552261\|20 | ···················· | ···················· | ····················· | ···················· | ···················· | ···················· |
| J | 7219 hCoV-19/England/MILK-994C8C/2020\|EPI_ISL_552263\|20 | ···················· | ···················· | ····················· | ···················· | ···················· | ···················· |
| J | 7220 hCoV-19/England/MILK-994C22/2020\|EPI_ISL_552264\|20 | ···················· | ···················· | ····················· | ···················· | ···················· | ···················· |
| J | 7221 hCoV-19/England/MILK-994DC5/2020\|EPI_ISL_552265\|20 | ···················· | ···················· | ····················· | ···················· | ···················· | ···················· |
| J | 7222 hCoV-19/England/MILK-99540B/2020\|EPI_ISL_552266\|20 | ···················· | ···················· | ····················· | ···················· | ···················· | ···················· |
| J | 7223 hCoV-19/England/MILK-994F74/2020\|EPI_ISL_552267\|20 | ···················· | ···················· | ····················· | ···················· | ···················· | ···················· |
| J | 7224 hCoV-19/England/ALDP-95A587/2020\|EPI_ISL_553508\|20 | ···················· | ···················· | ····················· | ···················· | ···················· | ···················· |
| J | 7225 hCoV-19/England/ALDP-95A842/2020\|EPI_ISL_553509\|20 | ···················· | ···················· | ····················· | ···················· | ···················· | ···················· |
| J | 7226 hCoV-19/England/ALDP-94D4B7/2020\|EPI_ISL_553522\|20 | ···················· | ···················· | ····················· | ···················· | ···················· | ···················· |
| J | 7227 hCoV-19/England/ALDP-95CB56/2020\|EPI_ISL_553523\|20 | ···················· | ···················· | ····················· | ···················· | ···················· | ···················· |
| J | 7228 hCoV-19/England/ALDP-95A44E/2020\|EPI_ISL_553524\|20 | ···················· | ···················· | ····················· | ···················· | ···················· | ···················· |
| J | 7229 hCoV-19/England/ALDP-955328/2020\|EPI_ISL_553525\|20 | ···················· | ···················· | ····················· | ···················· | ···················· | ···················· |
| J | 7230 hCoV-19/England/ALDP-9550E5/2020\|EPI_ISL_553526\|20 | ···················· | ···················· | ····················· | ···················· | ···················· | ···················· |
| J | 7231 hCoV-19/England/ALDP-9551A6/2020\|EPI_ISL_553528\|20 | ···················· | ···················· | ····················· | ···················· | ···················· | ···················· |
| J | 7232 hCoV-19/England/ALDP-95A01D/2020\|EPI_ISL_553529\|20 | ···················· | ···················· | ····················· | ···················· | ···················· | ···················· |
| J | 7233 hCoV-19/England/ALDP-95D241/2020\|EPI_ISL_553531\|20 | ···················· | ···················· | ····················· | ···················· | ···················· | ···················· |
| J | 7234 hCoV-19/England/ALDP-9552A3/2020\|EPI_ISL_553532\|20 | ···················· | ···················· | ····················· | ···················· | ···················· | ···················· |
| J | 7235 hCoV-19/England/ALDP-95D153/2020\|EPI_ISL_553533\|20 | ···················· | ···················· | ····················· | ···················· | ···················· | ···················· |
| J | 7236 hCoV-19/England/ALDP-959FA8/2020\|EPI_ISL_553534\|20 | ···················· | ···················· | ····················· | ···················· | ···················· | ···················· |
| J | 7237 hCoV-19/England/ALDP-95C96B/2020\|EPI_ISL_553535\|20 | ···················· | ···················· | ····················· | ···················· | ···················· | ···················· |
| J | 7238 hCoV-19/England/ALDP-94D4C6/2020\|EPI_ISL_553536\|20 | ···················· | ···················· | ····················· | ···················· | ···················· | ···················· |
| J | 7239 hCoV-19/England/ALDP-954DC1/2020\|EPI_ISL_553537\|20 | ···················· | ···················· | ····················· | ···················· | ···················· | ···················· |
| J | 7240 hCoV-19/England/ALDP-955531/2020\|EPI_ISL_553538\|20 | ···················· | ···················· | ····················· | ···················· | ···················· | ···················· |
| J | 7241 hCoV-19/England/ALDP-954E64/2020\|EPI_ISL_553540\|20 | ···················· | ···················· | ····················· | ···················· | ···················· | ···················· |
| J | 7242 hCoV-19/England/ALDP-95559B/2020\|EPI_ISL_553543\|20 | ···················· | ···················· | ····················· | ···················· | ···················· | ···················· |
| J | 7243 hCoV-19/England/ALDP-9550F4/2020\|EPI_ISL_553544\|20 | ···················· | ···················· | ····················· | ···················· | ···················· | ···················· |
| J | 7244 hCoV-19/England/ALDP-9552C1/2020\|EPI_ISL_553545\|20 | ···················· | ···················· | ····················· | ···················· | ···················· | ···················· |
| J | 7245 hCoV-19/England/ALDP-959FF3/2020\|EPI_ISL_553546\|20 | ···················· | ···················· | ····················· | ···················· | ···················· | ···················· |
| J | 7246 hCoV-19/England/ALDP-95D065/2020\|EPI_ISL_553547\|20 | ···················· | ···················· | ····················· | ···················· | ···················· | ···················· |
| J | 7247 hCoV-19/England/ALDP-95A4B7/2020\|EPI_ISL_553549\|20 | ···················· | ···················· | ····················· | ···················· | ···················· | ···················· |
| J | 7248 hCoV-19/England/ALDP-95C9D4/2020\|EPI_ISL_553551\|20 | ···················· | ···················· | ····················· | ···················· | ···················· | ···················· |
| J | 7249 hCoV-19/England/ALDP-95CB38/2020\|EPI_ISL_553552\|20 | ···················· | ···················· | ····················· | ···················· | ···················· | ···················· |
| J | 7250 hCoV-19/England/ALDP-955319/2020\|EPI_ISL_553553\|20 | ···················· | ···················· | ····················· | ···················· | ···················· | ···················· |
| J | 7251 hCoV-19/England/ALDP-95A45D/2020\|EPI_ISL_553554\|20 | ···················· | ···················· | ····················· | ···················· | ···················· | ···················· |
| J | 7252 hCoV-19/England/ALDP-95D0DE/2020\|EPI_ISL_553556\|20 | ···················· | ···················· | ····················· | ···················· | ···················· | ···················· |
| J | 7253 hCoV-19/England/ALDP-955021/2020\|EPI_ISL_553557\|20 | ···················· | ···················· | ····················· | ···················· | ···················· | ···················· |
| J | 7254 hCoV-19/England/ALDP-95570E/2020\|EPI_ISL_553558\|20 | ···················· | ···················· | ····················· | ···················· | ···················· | ···················· |
| J | 7255 hCoV-19/England/ALDP-95A46C/2020\|EPI_ISL_553559\|20 | ···················· | ···················· | ····················· | ···················· | ···················· | ···················· |
| J | 7256 hCoV-19/England/ALDP-95A745/2020\|EPI_ISL_553560\|20 | ···················· | ···················· | ····················· | ···················· | ···················· | ···················· |
| J | 7257 hCoV-19/England/ALDP-95CAE0/2020\|EPI_ISL_553561\|20 | ···················· | ···················· | ····················· | ···················· | ···················· | ···················· |
| J | 7258 hCoV-19/England/ALDP-954E0A/2020\|EPI_ISL_553562\|20 | ···················· | ···················· | ····················· | ···················· | ···················· | ···················· |
| J | 7259 hCoV-19/Wales/ALDP-94D693/2020\|EPI_ISL_553563\|2020 | ···················· | ···················· | ····················· | ···················· | ···················· | ···················· |
| J | 7260 hCoV-19/England/ALDP-95A402/2020\|EPI_ISL_553564\|20 | ···················· | ···················· | ····················· | ···················· | ···················· | ···················· |
| J | 7261 hCoV-19/England/ALDP-95D0CF/2020\|EPI_ISL_553566\|20 | ···················· | ···················· | ····················· | ···················· | ···················· | ···················· |
| J | 7262 hCoV-19/England/ALDP-9553CE/2020\|EPI_ISL_553569\|20 | ···················· | ···················· | ····················· | ···················· | ···················· | ···················· |
| J | 7263 hCoV-19/England/ALDP-95CB92/2020\|EPI_ISL_553570\|20 | ···················· | ···················· | ····················· | ···················· | ···················· | ···················· |
| J | 7264 hCoV-19/England/ALDP-954990/2020\|EPI_ISL_553571\|20 | ···················· | ···················· | ····················· | ···················· | ···················· | ···················· |
| J | 7265 hCoV-19/England/ALDP-95A6B1/2020\|EPI_ISL_553572\|20 | ···················· | ···················· | ····················· | ···················· | ···················· | ···················· |
| J | 7266 hCoV-19/England/ALDP-95A499/2020\|EPI_ISL_553573\|20 | ···················· | ···················· | ····················· | ···················· | ···················· | ···················· |
| J | 7267 hCoV-19/England/ALDP-95A420/2020\|EPI_ISL_553575\|20 | ···················· | ···················· | ····················· | ···················· | ···················· | ···················· |
| J | 7268 hCoV-19/England/ALDP-95A921/2020\|EPI_ISL_553576\|20 | ···················· | ···················· | ····················· | ···················· | ···················· | ···················· |
| J | 7269 hCoV-19/England/ALDP-954A24/2020\|EPI_ISL_553577\|20 | ···················· | ···················· | ····················· | ···················· | ···················· | ···················· |
| J | 7270 hCoV-19/England/ALDP-959FC6/2020\|EPI_ISL_553578\|20 | ···················· | ···················· | ····················· | ···················· | ···················· | ···················· |
| J | 7271 hCoV-19/England/ALDP-95D074/2020\|EPI_ISL_553579\|20 | ···················· | ···················· | ····················· | ···················· | ···················· | ···················· |
| J | 7272 hCoV-19/England/ALDP-95CB65/2020\|EPI_ISL_553580\|20 | ···················· | ···················· | ····················· | ···················· | ···················· | ···················· |
| J | 7273 hCoV-19/England/ALDP-9549AF/2020\|EPI_ISL_553581\|20 | ···················· | ···················· | ····················· | ···················· | ···················· | ···················· |
| J | 7274 hCoV-19/England/ALDP-95A43F/2020\|EPI_ISL_553582\|20 | ···················· | ···················· | ····················· | ···················· | ···················· | ···················· |

| | | | | | | | |
|---|---|---|---|---|---|---|---|
| J | 7275 hCoV-19/Wales/ALDP-955382/2020\|EPI_ISL_553583\|2020 | ···················· | ···················· | ····················· | ···················· | ···················· | ···················· |
| J | 7276 hCoV-19/England/ALDP-955197/2020\|EPI_ISL_553585\|20 | ···················· | ···················· | ····················· | ···················· | ···················· | ···················· |
| J | 7277 hCoV-19/England/ALDP-9552B2/2020\|EPI_ISL_553586\|20 | ···················· | ···················· | ····················· | ···················· | ···················· | ···················· |
| J | 7278 hCoV-19/England/ALDP-95A5A5/2020\|EPI_ISL_553587\|20 | ···················· | ···················· | ····················· | ···················· | ···················· | ···················· |
| J | 7279 hCoV-19/England/ALDP-954FAD/2020\|EPI_ISL_553588\|20 | ···················· | ···················· | ····················· | ···················· | ···················· | ···················· |
| J | 7280 hCoV-19/England/ALDP-95A208/2020\|EPI_ISL_553589\|20 | ···················· | ···················· | ····················· | ···················· | ···················· | ···················· |
| J | 7281 hCoV-19/England/ALDP-954E37/2020\|EPI_ISL_553590\|20 | ···················· | ···················· | ····················· | ···················· | ···················· | ···················· |
| J | 7282 hCoV-19/England/ALDP-95509A/2020\|EPI_ISL_553591\|20 | ···················· | ···················· | ····················· | ···················· | ···················· | ···················· |
| J | 7283 hCoV-19/England/ALDP-95CBA1/2020\|EPI_ISL_553592\|20 | ···················· | ···················· | ····················· | ···················· | ···················· | ···················· |
| J | 7284 hCoV-19/England/ALDP-954CF1/2020\|EPI_ISL_553593\|20 | ···················· | ···················· | ····················· | ···················· | ···················· | ···················· |
| J | 7285 hCoV-19/England/ALDP-9551E2/2020\|EPI_ISL_553594\|20 | ···················· | ···················· | ····················· | ···················· | ···················· | ···················· |
| J | 7286 hCoV-19/England/ALDP-954E55/2020\|EPI_ISL_553595\|20 | ···················· | ···················· | ····················· | ···················· | ···················· | ···················· |
| J | 7287 hCoV-19/England/ALDP-95508B/2020\|EPI_ISL_553596\|20 | ···················· | ···················· | ····················· | ···················· | ···················· | ···················· |
| J | 7288 hCoV-19/England/ALDP-955294/2020\|EPI_ISL_553597\|20 | ···················· | ···················· | ····················· | ···················· | ···················· | ···················· |
| J | 7289 hCoV-19/England/ALDP-95562F/2020\|EPI_ISL_553598\|20 | ···················· | ···················· | ····················· | ···················· | ···················· | ···················· |
| J | 7290 hCoV-19/England/MILK-9583CB/2020\|EPI_ISL_554801\|20 | ···················· | ···················· | ····················· | ···················· | ···················· | ···················· |
| J | 7291 hCoV-19/England/ALDP-944007/2020\|EPI_ISL_554802\|20 | ···················· | ·A·················· | ····················· | ···················· | ···················· | ···················· |
| J | 7292 hCoV-19/England/ALDP-944641/2020\|EPI_ISL_554804\|20 | ···················· | ···················· | ····················· | ···················· | ···················· | ···················· |
| J | 7293 hCoV-19/England/MILK-958000/2020\|EPI_ISL_554805\|20 | ···················· | ···················· | ····················· | ···················· | ···················· | ···················· |
| J | 7294 hCoV-19/England/MILK-958097/2020\|EPI_ISL_554806\|20 | ···················· | ···················· | ····················· | ···················· | ···················· | ···················· |
| J | 7295 hCoV-19/England/ALDP-94423E/2020\|EPI_ISL_554808\|20 | ···················· | ···················· | ····················· | ···················· | ···················· | ···················· |
| J | 7296 hCoV-19/England/ALDP-94424D/2020\|EPI_ISL_554809\|20 | ···················· | ···················· | ····················· | ···················· | ···················· | ···················· |
| J | 7297 hCoV-19/England/ALDP-9445CC/2020\|EPI_ISL_554810\|20 | ···················· | ···················· | ····················· | ···················· | ···················· | ···················· |
| J | 7298 hCoV-19/England/ALDP-9441D7/2020\|EPI_ISL_554811\|20 | ···················· | ···················· | ····················· | ···················· | ···················· | ···················· |
| J | 7299 hCoV-19/England/ALDP-9441AA/2020\|EPI_ISL_554812\|20 | ···················· | ···················· | ····················· | ···················· | ···················· | ···················· |
| J | 7300 hCoV-19/England/ALDP-9443E0/2020\|EPI_ISL_554813\|20 | ···················· | ···················· | ····················· | ···················· | ···················· | ···················· |
| J | 7301 hCoV-19/England/MILK-958589/2020\|EPI_ISL_554814\|20 | ···················· | ···················· | ····················· | ···················· | ···················· | ···················· |
| J | 7302 hCoV-19/England/ALDP-94418C/2020\|EPI_ISL_554815\|20 | ···················· | ···················· | ····················· | ···················· | ···················· | ···················· |
| J | 7303 hCoV-19/England/ALDP-94408F/2020\|EPI_ISL_554816\|20 | ···················· | ···················· | ····················· | ···················· | ···················· | ···················· |
| J | 7304 hCoV-19/England/MILK-95997D/2020\|EPI_ISL_554817\|20 | ···················· | ···················· | ····················· | ···················· | ···················· | ···················· |
| J | 7305 hCoV-19/England/MILK-957FD7/2020\|EPI_ISL_554818\|20 | ···················· | ···················· | ····················· | ···················· | ···················· | ···················· |
| J | 7306 hCoV-19/England/MILK-958079/2020\|EPI_ISL_554819\|20 | ···················· | ···················· | ····················· | ···················· | ···················· | ···················· |
| J | 7307 hCoV-19/England/ALDP-943F56/2020\|EPI_ISL_554820\|20 | ···················· | ···················· | ····················· | ···················· | ···················· | ···················· |
| J | 7308 hCoV-19/England/MILK-9583BC/2020\|EPI_ISL_554821\|20 | ···················· | ···················· | ····················· | ···················· | ···················· | ···················· |
| J | 7309 hCoV-19/England/MILK-958598/2020\|EPI_ISL_554822\|20 | ···················· | ···················· | ····················· | ···················· | ···················· | ···················· |
| J | 7310 hCoV-19/England/MILK-9580D3/2020\|EPI_ISL_554824\|20 | ···················· | ···················· | ····················· | ···················· | ···················· | ···················· |
| J | 7311 hCoV-19/England/MILK-958A3F/2020\|EPI_ISL_554825\|20 | ···················· | ···················· | ····················· | ···················· | ···················· | ···················· |
| J | 7312 hCoV-19/England/MILK-957C94/2020\|EPI_ISL_554826\|20 | ···················· | ···················· | ····················· | ···················· | ···················· | ···················· |
| J | 7313 hCoV-19/England/MILK-95854D/2020\|EPI_ISL_554828\|20 | ···················· | ···················· | ····················· | ···················· | ···················· | ···················· |
| J | 7314 hCoV-19/England/MILK-957C67/2020\|EPI_ISL_554829\|20 | ···················· | ···················· | ····················· | ···················· | ···················· | ···················· |
| J | 7315 hCoV-19/England/ALDP-943F47/2020\|EPI_ISL_554830\|20 | ···················· | ···················· | ····················· | ···················· | ···················· | ···················· |
| J | 7316 hCoV-19/England/MILK-958677/2020\|EPI_ISL_554839\|20 | ···················· | ···················· | ····················· | ···················· | ···················· | ···················· |
| J | 7317 hCoV-19/England/ALDP-94427A/2020\|EPI_ISL_554841\|20 | ···················· | ···················· | ····················· | ···················· | ···················· | ···················· |
| J | 7318 hCoV-19/England/ALDP-9442D4/2020\|EPI_ISL_554853\|20 | ···················· | ···················· | ····················· | ···················· | ···················· | ···················· |
| J | 7319 hCoV-19/England/MILK-9585B6/2020\|EPI_ISL_554854\|20 | ···················· | ···················· | ····················· | ···················· | ···················· | ···················· |
| J | 7320 hCoV-19/England/MILK-958747/2020\|EPI_ISL_554855\|20 | ···················· | ···················· | ····················· | ···················· | ···················· | ···················· |
| J | 7321 hCoV-19/England/MILK-958A6C/2020\|EPI_ISL_554857\|20 | ···················· | ···················· | ····················· | ···················· | ···················· | ···················· |
| J | 7322 hCoV-19/England/MILK-958B4B/2020\|EPI_ISL_554860\|20 | ···················· | ···················· | ····················· | ···················· | ···················· | ···················· |
| J | 7323 hCoV-19/England/ALDP-959E41/2020\|EPI_ISL_554861\|20 | ···················· | ···················· | ····················· | ···················· | ···················· | ···················· |
| J | 7324 hCoV-19/England/ALDP-95CE6C/2020\|EPI_ISL_554862\|20 | ···················· | ···················· | ····················· | ···················· | ···················· | ···················· |
| J | 7325 hCoV-19/England/ALDP-95C28E/2020\|EPI_ISL_554863\|20 | ···················· | ···················· | ····················· | ···················· | ···················· | ···················· |
| J | 7326 hCoV-19/England/ALDP-94EA5A/2020\|EPI_ISL_554864\|20 | ···················· | ···················· | ····················· | ···················· | ···················· | ···················· |
| J | 7327 hCoV-19/England/ALDP-94E9A8/2020\|EPI_ISL_554865\|20 | ···················· | ···················· | ····················· | ···················· | ···················· | ···················· |
| J | 7328 hCoV-19/England/ALDP-95C7DA/2020\|EPI_ISL_554866\|20 | ···················· | ···················· | ····················· | ···················· | ···················· | ···················· |
| J | 7329 hCoV-19/England/ALDP-95AAB5/2020\|EPI_ISL_554867\|20 | ···················· | ···················· | ····················· | ···················· | ···················· | ···················· |
| J | 7330 hCoV-19/England/ALDP-94EA4B/2020\|EPI_ISL_554870\|20 | ···················· | ···················· | ····················· | ···················· | ···················· | ···················· |
| J | 7331 hCoV-19/England/ALDP-95CD9C/2020\|EPI_ISL_554871\|20 | ···················· | ···················· | ····················· | ···················· | ···················· | ···················· |
| J | 7332 hCoV-19/England/ALDP-94D842/2020\|EPI_ISL_554872\|20 | ···················· | ···················· | ····················· | ···················· | ···················· | ···················· |
| J | 7333 hCoV-19/England/ALDP-95CD41/2020\|EPI_ISL_554873\|20 | ···················· | ···················· | ····················· | ···················· | ···················· | ···················· |
| J | 7334 hCoV-19/England/ALDP-95CE02/2020\|EPI_ISL_554874\|20 | ···················· | ···················· | ····················· | ···················· | ···················· | ···················· |
| J | 7335 hCoV-19/England/ALDP-95CC35/2020\|EPI_ISL_554875\|20 | ···················· | ···················· | ····················· | ···················· | ···················· | ···················· |
| J | 7336 hCoV-19/England/ALDP-95C8B9/2020\|EPI_ISL_554876\|20 | ···················· | ···················· | ····················· | ···················· | ···················· | ···················· |
| J | 7337 hCoV-19/England/ALDP-95CFB4/2020\|EPI_ISL_554877\|20 | ···················· | ···················· | ····················· | ···················· | ···················· | ···················· |
| J | 7338 hCoV-19/England/ALDP-94D54B/2020\|EPI_ISL_554878\|20 | ···················· | ···················· | ····················· | ···················· | ···················· | ···················· |
| J | 7339 hCoV-19/England/ALDP-94E86F/2020\|EPI_ISL_554879\|20 | ···················· | ···················· | ····················· | ···················· | ···················· | ···················· |
| J | 7340 hCoV-19/England/ALDP-95CE20/2020\|EPI_ISL_554880\|20 | ···················· | ···················· | ····················· | ···················· | ···················· | ···················· |
| J | 7341 hCoV-19/England/ALDP-95A754/2020\|EPI_ISL_554881\|20 | ···················· | ···················· | ····················· | ···················· | ···················· | ···················· |
| J | 7342 hCoV-19/England/ALDP-95C29D/2020\|EPI_ISL_554883\|20 | ···················· | ···················· | ····················· | ···················· | ···················· | ···················· |
| J | 7343 hCoV-19/England/ALDP-95CEA8/2020\|EPI_ISL_554884\|20 | ···················· | ···················· | ····················· | ···················· | ···················· | ···················· |
| J | 7344 hCoV-19/England/ALDP-95C60A/2020\|EPI_ISL_554885\|20 | ···················· | ···················· | ····················· | ···················· | ···················· | ···················· |
| J | 7345 hCoV-19/England/ALDP-95C79E/2020\|EPI_ISL_554888\|20 | ···················· | ···················· | ····················· | ···················· | ···················· | ···················· |
| J | 7346 hCoV-19/England/ALDP-95C813/2020\|EPI_ISL_554889\|20 | ···················· | ···················· | ····················· | ···················· | ···················· | ···················· |
| J | 7347 hCoV-19/England/ALDP-95C084/2020\|EPI_ISL_554890\|20 | ···················· | ···················· | ····················· | ···················· | ···················· | ···················· |
| J | 7348 hCoV-19/Wales/ALDP-95C0A2/2020\|EPI_ISL_554891\|2020 | ···················· | ···················· | ····················· | ···················· | ···················· | ···················· |
| J | 7349 hCoV-19/England/ALDP-95A781/2020\|EPI_ISL_554892\|20 | ···················· | ···················· | ····················· | ···················· | ···················· | ···················· |
| J | 7350 hCoV-19/England/ALDP-95AC73/2020\|EPI_ISL_554893\|20 | ···················· | ···················· | ····················· | ···················· | ···················· | ···················· |
| J | 7351 hCoV-19/England/ALDP-95A97C/2020\|EPI_ISL_554894\|20 | ···················· | ···················· | ····················· | ···················· | ···················· | ···················· |
| J | 7352 hCoV-19/England/ALDP-95C172/2020\|EPI_ISL_554895\|20 | ···················· | ···················· | ····················· | ···················· | ···················· | ···················· |
| J | 7353 hCoV-19/England/ALDP-94D745/2020\|EPI_ISL_554896\|20 | ···················· | ···················· | ····················· | ···················· | ···················· | ···················· |
| J | 7354 hCoV-19/England/ALDP-95CD50/2020\|EPI_ISL_554899\|20 | ···················· | ···················· | ····················· | ···················· | ···················· | ···················· |

| | | | | | | | | |
|---|---|---|---|---|---|---|---|---|
| J | 7355 | hCoV-19/England/ALDP-8B9F4D/2020\|EPI_ISL_557041\|20 | ···················· | ···················· | ···················· | ···················· | ···················· | ···················· |
| J | 7356 | hCoV-19/England/ALDP-8E02DB/2020\|EPI_ISL_557044\|20 | ···················· | ···················· | ···················· | ···················· | ···················· | ···················· |
| J | 7357 | hCoV-19/England/ALDP-8D74A4/2020\|EPI_ISL_557045\|20 | ···················· | ···················· | ···················· | ···················· | ···················· | ···················· |
| J | 7358 | hCoV-19/England/ALDP-8DA650/2020\|EPI_ISL_557046\|20 | ···················· | ···················· | ···················· | ···················· | ···················· | ···················· |
| J | 7359 | hCoV-19/England/ALDP-8D1774/2020\|EPI_ISL_557047\|20 | ···················· | ···················· | ···················· | ···················· | ···················· | ···················· |
| J | 7360 | hCoV-19/England/ALDP-8B9903/2020\|EPI_ISL_557049\|20 | ···················· | ···················· | ···················· | ···················· | ···················· | ···················· |
| J | 7361 | hCoV-19/England/ALDP-90FCE6/2020\|EPI_ISL_557050\|20 | ···················· | ···················· | ···················· | ···················· | ···················· | ···················· |
| J | 7362 | hCoV-19/England/ALDP-8DD09B/2020\|EPI_ISL_557051\|20 | ···················· | ···················· | ···················· | ···················· | ···················· | ···················· |
| J | 7363 | hCoV-19/England/ALDP-8E1762/2020\|EPI_ISL_557052\|20 | ···················· | ···················· | ···················· | ···················· | ···················· | ···················· |
| J | 7364 | hCoV-19/England/ALDP-8E2567/2020\|EPI_ISL_557053\|20 | ···················· | ···················· | ···················· | ···················· | ···················· | ···················· |
| J | 7365 | hCoV-19/England/ALDP-8DBA9F/2020\|EPI_ISL_557054\|20 | ···················· | ···················· | ···················· | ···················· | ···················· | ···················· |
| J | 7366 | hCoV-19/England/ALDP-8E2F2D/2020\|EPI_ISL_557055\|20 | ···················· | ···················· | ···················· | ···················· | ···················· | ···················· |
| J | 7367 | hCoV-19/England/ALDP-8FC8CB/2020\|EPI_ISL_557056\|20 | ···················· | ···················· | ···················· | ···················· | ···················· | ···················· |
| J | 7368 | hCoV-19/England/ALDP-8DFFD8/2020\|EPI_ISL_557057\|20 | ···················· | ···················· | ···················· | ···················· | ···················· | ···················· |
| J | 7369 | hCoV-19/England/ALDP-8D41CE/2020\|EPI_ISL_557058\|20 | ···················· | ···················· | ···················· | ···················· | ···················· | ···················· |
| J | 7370 | hCoV-19/England/ALDP-8DCD0E/2020\|EPI_ISL_557059\|20 | ···················· | ···················· | ···················· | ···················· | ···················· | ···················· |
| J | 7371 | hCoV-19/England/ALDP-8DDEFB/2020\|EPI_ISL_557060\|20 | ···················· | ···················· | ···················· | ···················· | ···················· | ···················· |
| J | 7372 | hCoV-19/England/ALDP-901437/2020\|EPI_ISL_557061\|20 | ···················· | ···················· | ···················· | ···················· | ···················· | ···················· |
| J | 7373 | hCoV-19/England/ALDP-8FEF3D/2020\|EPI_ISL_557062\|20 | ···················· | ···················· | ···················· | ···················· | ···················· | ···················· |
| J | 7374 | hCoV-19/England/ALDP-8FF2E8/2020\|EPI_ISL_557063\|20 | ···················· | ···················· | ···················· | ···················· | ···················· | ···················· |
| J | 7375 | hCoV-19/England/ALDP-8DF873/2020\|EPI_ISL_557064\|20 | ···················· | ···················· | ···················· | ···················· | ···················· | ···················· |
| J | 7376 | hCoV-19/England/ALDP-8FCA7A/2020\|EPI_ISL_557065\|20 | ···················· | ···················· | ···················· | ···················· | ···················· | ···················· |
| J | 7377 | hCoV-19/England/ALDP-8DE30A/2020\|EPI_ISL_557066\|20 | ···················· | ···················· | ···················· | ···················· | ···················· | ···················· |
| J | 7378 | hCoV-19/England/ALDP-8DB75C/2020\|EPI_ISL_557067\|20 | ···················· | ···················· | ···················· | ···················· | ···················· | ···················· |
| J | 7379 | hCoV-19/England/ALDP-8D7125/2020\|EPI_ISL_557068\|20 | ···················· | ···················· | ···················· | ···················· | ···················· | ···················· |
| J | 7380 | hCoV-19/England/ALDP-9011E5/2020\|EPI_ISL_557069\|20 | ···················· | ···················· | ···················· | ···················· | ···················· | ···················· |
| J | 7381 | hCoV-19/England/ALDP-8D453B/2020\|EPI_ISL_557070\|20 | ···················· | ···················· | ···················· | ···················· | ···················· | ···················· |
| J | 7382 | hCoV-19/England/ALDP-8B672A/2020\|EPI_ISL_557072\|20 | ···················· | ···················· | ···················· | ···················· | ···················· | ···················· |
| J | 7383 | hCoV-19/England/ALDP-8D634E/2020\|EPI_ISL_557073\|20 | ···················· | ···················· | ···················· | ···················· | ···················· | ···················· |
| J | 7384 | hCoV-19/England/ALDP-8B6283/2020\|EPI_ISL_557075\|20 | ···················· | ···················· | ···················· | ···················· | ···················· | ···················· |
| J | 7385 | hCoV-19/England/ALDP-8DE1C4/2020\|EPI_ISL_557076\|20 | ···················· | ···················· | ···················· | ···················· | ···················· | ···················· |
| J | 7386 | hCoV-19/England/ALDP-8CF454/2020\|EPI_ISL_557078\|20 | ···················· | ···················· | ···················· | ···················· | ···················· | ···················· |
| J | 7387 | hCoV-19/England/ALDP-8E1814/2020\|EPI_ISL_557080\|20 | ···················· | ···················· | ···················· | ···················· | ···················· | ···················· |
| J | 7388 | hCoV-19/England/ALDP-8E388B/2020\|EPI_ISL_557082\|20 | ···················· | ···················· | ···················· | ···················· | ···················· | ···················· |
| J | 7389 | hCoV-19/England/ALDP-8E2E8A/2020\|EPI_ISL_557083\|20 | ···················· | ···················· | ···················· | ···················· | ···················· | ···················· |
| J | 7390 | hCoV-19/England/ALDP-8CEE75/2020\|EPI_ISL_557084\|20 | ···················· | ···················· | ···················· | ···················· | ···················· | ···················· |
| J | 7391 | hCoV-19/England/ALDP-8CF83A/2020\|EPI_ISL_557086\|20 | ···················· | ···················· | ···················· | ···················· | ···················· | ···················· |
| J | 7392 | hCoV-19/England/ALDP-8CED1E/2020\|EPI_ISL_557088\|20 | ···················· | ···················· | ···················· | ···················· | ···················· | ···················· |
| J | 7393 | hCoV-19/Wales/ALDP-8D8D3F/2020\|EPI_ISL_557089\|2020 | ···················· | ···················· | ···················· | ···················· | ···················· | ···················· |
| J | 7394 | hCoV-19/England/ALDP-8D1E9D/2020\|EPI_ISL_557090\|20 | ···················· | ···················· | ···················· | ···················· | ···················· | ···················· |
| J | 7395 | hCoV-19/England/ALDP-8B5D83/2020\|EPI_ISL_557091\|20 | ···················· | ···················· | ···················· | ···················· | ···················· | ···················· |
| J | 7396 | hCoV-19/England/ALDP-8D5646/2020\|EPI_ISL_557092\|20 | ···················· | ···················· | ···················· | ···················· | ···················· | ···················· |
| J | 7397 | hCoV-19/England/ALDP-9015CB/2020\|EPI_ISL_557093\|20 | ···················· | ···················· | ···················· | ···················· | ···················· | ···················· |
| J | 7398 | hCoV-19/England/ALDP-8E2B29/2020\|EPI_ISL_557095\|20 | ···················· | ···················· | ···················· | ···················· | ···················· | ···················· |
| J | 7399 | hCoV-19/England/ALDP-8E2B65/2020\|EPI_ISL_557097\|20 | ···················· | ···················· | ···················· | ···················· | ···················· | ···················· |
| J | 7400 | hCoV-19/England/ALDP-8B6F9B/2020\|EPI_ISL_557098\|20 | ···················· | ···················· | ···················· | ···················· | ···················· | ···················· |
| J | 7401 | hCoV-19/England/ALDP-8B9842/2020\|EPI_ISL_557099\|20 | ···················· | ···················· | ···················· | ···················· | ···················· | ···················· |
| J | 7402 | hCoV-19/England/MILK-562D6A/2020\|EPI_ISL_558372\|20 | ···················· | ···················· | ···················· | ···················· | ···················· | ···················· |
| J | 7403 | hCoV-19/England/MILK-56C53B/2020\|EPI_ISL_558374\|20 | ···················· | ···················· | ···················· | ···················· | ···················· | ···················· |
| J | 7404 | hCoV-19/England/MILK-56259F/2020\|EPI_ISL_558375\|20 | ···················· | ···················· | ···················· | ···················· | ···················· | ···················· |
| J | 7405 | hCoV-19/England/MILK-592763/2020\|EPI_ISL_558376\|20 | ···················· | ···················· | ···················· | ···················· | ···················· | ···················· |
| J | 7406 | hCoV-19/England/MILK-55E790/2020\|EPI_ISL_558378\|20 | ···················· | ···················· | ···················· | ···················· | ···················· | ···················· |
| J | 7407 | hCoV-19/England/ALDP-59977B/2020\|EPI_ISL_558379\|20 | ···················· | ···················· | ···················· | ···················· | ···················· | ···················· |
| J | 7408 | hCoV-19/England/MILK-56B89D/2020\|EPI_ISL_558380\|20 | ···················· | ···················· | ···················· | ···················· | ···················· | ···················· |
| J | 7409 | hCoV-19/England/MILK-568CD0/2020\|EPI_ISL_558381\|20 | ···················· | ···················· | ···················· | ···················· | ···················· | ···················· |
| J | 7410 | hCoV-19/England/MILK-57A163/2020\|EPI_ISL_558382\|20 | ···················· | ···················· | ···················· | ···················· | ········T··········· | ···················· |
| J | 7411 | hCoV-19/England/MILK-5784E4/2020\|EPI_ISL_558383\|20 | ···················· | ···················· | ···················· | ···················· | ···················· | ···················· |
| J | 7412 | hCoV-19/England/MILK-55F322/2020\|EPI_ISL_558384\|20 | ··········T········· | ···················· | ···················· | ···················· | ···················· | ···················· |
| J | 7413 | hCoV-19/England/MILK-568D0A/2020\|EPI_ISL_558385\|20 | ···················· | ···················· | ···················· | ···················· | ···················· | ···················· |
| J | 7414 | hCoV-19/England/MILK-56BFD4/2020\|EPI_ISL_558386\|20 | ···················· | ···················· | ···················· | ···················· | ···················· | ···················· |
| J | 7415 | hCoV-19/England/MILK-57314C/2020\|EPI_ISL_558388\|20 | ···················· | ···················· | ···················· | ···················· | ···················· | ···················· |
| J | 7416 | hCoV-19/England/ALDP-59AAF9/2020\|EPI_ISL_558389\|20 | ···················· | ···················· | ···················· | ···················· | ···················· | ···················· |
| J | 7417 | hCoV-19/England/MILK-58F374/2020\|EPI_ISL_558390\|20 | ···················· | ···················· | ···················· | ···················· | ···················· | ···················· |
| J | 7418 | hCoV-19/England/MILK-559F15/2020\|EPI_ISL_558391\|20 | ···················· | ···················· | ···················· | ···················· | ···················· | ···················· |
| J | 7419 | hCoV-19/England/MILK-559DB1/2020\|EPI_ISL_558393\|20 | ···················· | ···················· | ···················· | ···················· | ···················· | ···················· |
| J | 7420 | hCoV-19/England/ALDP-59AB8D/2020\|EPI_ISL_558394\|20 | ···················· | ·A·················· | ···················· | ···················· | ···················· | ···················· |
| J | 7421 | hCoV-19/England/ALDP-59A8D1/2020\|EPI_ISL_558395\|20 | ···················· | ···················· | ···················· | ···················· | ···················· | ···················· |
| J | 7422 | hCoV-19/England/ALDP-59C4DB/2020\|EPI_ISL_558396\|20 | ···················· | ···················· | ···················· | ···················· | ···················· | ···················· |
| J | 7423 | hCoV-19/England/MILK-562E76/2020\|EPI_ISL_558397\|20 | ···················· | ···················· | ···················· | ···················· | ···················· | ···················· |
| J | 7424 | hCoV-19/England/MILK-562ABE/2020\|EPI_ISL_558399\|20 | ···················· | ···················· | ···················· | ···················· | ···················· | ···················· |
| J | 7425 | hCoV-19/England/MILK-9937F5/2020\|EPI_ISL_552026\|20 | ···················· | ···················· | ···················· | ···················· | ···················· | ···················· |
| J | 7426 | hCoV-19/England/MILK-992BAF/2020\|EPI_ISL_552027\|20 | ···················· | ·A·················· | ···················· | ···················· | ···················· | ···A················ |
| J | 7427 | hCoV-19/England/MILK-9927F6/2020\|EPI_ISL_552028\|20 | ···················· | ···················· | ···················· | ···················· | ···················· | ···················· |
| J | 7428 | hCoV-19/England/MILK-992E4C/2020\|EPI_ISL_552029\|20 | ···················· | ···················· | ···················· | ···················· | ···················· | ···················· |
| J | 7429 | hCoV-19/England/MILK-9933C4/2020\|EPI_ISL_552030\|20 | ···················· | ···················· | ···················· | ···················· | ···················· | ···················· |
| J | 7430 | hCoV-19/England/MILK-99342B/2020\|EPI_ISL_552031\|20 | ···················· | ···················· | ···················· | ···················· | ···················· | ···················· |
| J | 7431 | hCoV-19/England/MILK-993133/2020\|EPI_ISL_552032\|20 | ···················· | ···················· | ···················· | ···················· | ···················· | ···················· |
| J | 7432 | hCoV-19/England/MILK-992D8B/2020\|EPI_ISL_552033\|20 | ···················· | ···················· | ···················· | ···················· | ···················· | ···················· |
| J | 7433 | hCoV-19/England/MILK-9934A3/2020\|EPI_ISL_552034\|20 | ···················· | ···················· | ···················· | ···················· | ···················· | ··············A····· |
| J | 7434 | hCoV-19/England/MILK-99340D/2020\|EPI_ISL_552035\|20 | ···················· | ···················· | ···················· | ···················· | ···················· | ···················· |

| | | | | | | | | |
|---|---|---|---|---|---|---|---|---|
| J | 7435 | hCoV-19/England/MILK-992680/2020\|EPI_ISL_552036\|20 | ···················· | ·A·······A·········· | ····················· | ···················· | ···················· | ···················· |
| J | 7436 | hCoV-19/England/MILK-993027/2020\|EPI_ISL_552038\|20 | ···················· | ···················· | ····················· | ···················· | ···················· | ···················· |
| J | 7437 | hCoV-19/England/MILK-992D4F/2020\|EPI_ISL_552039\|20 | ···················· | ···················· | ····················· | ···················· | ···················· | ···················· |
| J | 7438 | hCoV-19/Wales/MILK-993476/2020\|EPI_ISL_552040\|2020 | ···················· | ···················· | ····················· | ···················· | ···················· | ···················· |
| J | 7439 | hCoV-19/England/MILK-9925ED/2020\|EPI_ISL_552041\|20 | ···················· | ···················· | ····················· | ···················· | ···················· | ···················· |
| J | 7440 | hCoV-19/England/MILK-991E2F/2020\|EPI_ISL_552042\|20 | ···················· | ···················· | ····················· | ···················· | ···················· | ···················· |
| J | 7441 | hCoV-19/England/MILK-993203/2020\|EPI_ISL_552043\|20 | ···················· | ···················· | ····················· | ···················· | ···················· | ···················· |
| J | 7442 | hCoV-19/Wales/MILK-992C51/2020\|EPI_ISL_552045\|2020 | ···················· | ···················· | ····················· | ···················· | ···················· | ···················· |
| J | 7443 | hCoV-19/England/MILK-9925CF/2020\|EPI_ISL_552047\|20 | ···················· | ···················· | ····················· | ···················· | ···················· | ···················· |
| J | 7444 | hCoV-19/England/MILK-993072/2020\|EPI_ISL_552048\|20 | ···················· | ···················· | ····················· | ···················· | ···················· | ···················· |
| J | 7445 | hCoV-19/England/MILK-992DA9/2020\|EPI_ISL_552049\|20 | ···················· | ···················· | ····················· | ···················· | ···················· | ···················· |
| J | 7446 | hCoV-19/England/MILK-9928E4/2020\|EPI_ISL_552050\|20 | ···················· | ···················· | ····················· | ···················· | ······T············· | ···················· |
| J | 7447 | hCoV-19/England/CAMC-99141E/2020\|EPI_ISL_552051\|20 | ···················· | ···················· | ····················· | ···················· | ···················· | ···················· |
| J | 7448 | hCoV-19/England/MILK-992CAC/2020\|EPI_ISL_552052\|20 | ···················· | ···················· | ····················· | ···················· | ···················· | ···················· |
| J | 7449 | hCoV-19/England/MILK-9930AF/2020\|EPI_ISL_552054\|20 | ···················· | ···················· | ····················· | ···················· | ···················· | ···················· |
| J | 7450 | hCoV-19/England/MILK-992D12/2020\|EPI_ISL_552055\|20 | ···················· | ···················· | ····················· | ···················· | ···················· | ···················· |
| J | 7451 | hCoV-19/Wales/MILK-9928A8/2020\|EPI_ISL_552056\|2020 | ···················· | ···················· | ····················· | ···················· | ···················· | ···················· |
| J | 7452 | hCoV-19/England/MILK-992626/2020\|EPI_ISL_552057\|20 | ···················· | ···················· | ····················· | ···················· | ···················· | ···················· |
| J | 7453 | hCoV-19/Wales/MILK-993115/2020\|EPI_ISL_552058\|2020 | ···················· | ···················· | ····················· | ···················· | ···················· | ···················· |
| J | 7454 | hCoV-19/England/MILK-992ADF/2020\|EPI_ISL_552059\|20 | ···················· | ···················· | ····················· | ···················· | ···················· | ···················· |
| J | 7455 | hCoV-19/England/MILK-99290F/2020\|EPI_ISL_552060\|20 | ···················· | ·A·················· | ····················· | ···················· | ···················· | ···················· |
| J | 7456 | hCoV-19/England/MILK-992C60/2020\|EPI_ISL_552061\|20 | ···················· | ·A·················· | ····················· | ···················· | ···················· | ···················· |
| J | 7457 | hCoV-19/Wales/MILK-992CE8/2020\|EPI_ISL_552062\|2020 | ···················· | ···················· | ····················· | ···················· | ···················· | ···················· |
| J | 7458 | hCoV-19/England/MILK-992635/2020\|EPI_ISL_552063\|20 | ···················· | ···················· | ····················· | ···················· | ···················· | ···················· |
| J | 7459 | hCoV-19/England/CAMC-991636/2020\|EPI_ISL_552065\|20 | ···················· | ···················· | ····················· | ···················· | ···················· | ···················· |
| J | 7460 | hCoV-19/Wales/MILK-9928D5/2020\|EPI_ISL_552066\|2020 | ···················· | ···················· | ····················· | ···················· | ···················· | ···················· |
| J | 7461 | hCoV-19/England/MILK-993090/2020\|EPI_ISL_552067\|20 | ···················· | ···················· | ····················· | ···················· | ···················· | ···················· |
| J | 7462 | hCoV-19/England/MILK-9937E6/2020\|EPI_ISL_552068\|20 | ···················· | ···················· | ····················· | ···················· | ······T············· | ···················· |
| J | 7463 | hCoV-19/England/MILK-993124/2020\|EPI_ISL_552069\|20 | ···················· | ···················· | ····················· | ···················· | ···················· | ···················· |
| J | 7464 | hCoV-19/England/MILK-992F85/2020\|EPI_ISL_552070\|20 | ···················· | ···················· | ····················· | ···················· | ···················· | ···················· |
| J | 7465 | hCoV-19/England/MILK-992EF1/2020\|EPI_ISL_552071\|20 | ···················· | ···················· | ····················· | ···················· | ···················· | ···················· |
| J | 7466 | hCoV-19/England/MILK-993045/2020\|EPI_ISL_552072\|20 | ···················· | ···················· | ····················· | ···················· | ···················· | ···················· |
| J | 7467 | hCoV-19/England/CAMC-991627/2020\|EPI_ISL_552073\|20 | ···················· | ···················· | ····················· | ···················· | ···················· | ···················· |
| J | 7468 | hCoV-19/England/MILK-9931D9/2020\|EPI_ISL_552075\|20 | ···················· | ···················· | ····················· | ···················· | ···················· | ···················· |
| J | 7469 | hCoV-19/England/MILK-9927AB/2020\|EPI_ISL_552076\|20 | ···················· | ···················· | ····················· | ···················· | ···················· | ···················· |
| J | 7470 | hCoV-19/England/MILK-9929A5/2020\|EPI_ISL_552077\|20 | ···················· | ···················· | ····················· | ···················· | ···················· | ···················· |
| J | 7471 | hCoV-19/England/MILK-993546/2020\|EPI_ISL_552078\|20 | ···················· | ···················· | ····················· | ···················· | ···················· | ···················· |
| J | 7472 | hCoV-19/England/MILK-992BDC/2020\|EPI_ISL_552079\|20 | ···················· | ···················· | ····················· | ···················· | ···················· | ···················· |
| J | 7473 | hCoV-19/England/MILK-99317F/2020\|EPI_ISL_552080\|20 | ···················· | ···················· | ····················· | ···················· | ···················· | ···················· |
| J | 7474 | hCoV-19/Wales/MILK-993467/2020\|EPI_ISL_552081\|2020 | ···················· | ···················· | ····················· | ···················· | ···················· | ···················· |
| J | 7475 | hCoV-19/England/MILK-992671/2020\|EPI_ISL_552082\|20 | ···················· | ···················· | ····················· | ···················· | ···················· | ···················· |
| J | 7476 | hCoV-19/England/MILK-9931F7/2020\|EPI_ISL_552084\|20 | ···················· | ···················· | ····················· | ···················· | ···················· | ···················· |
| J | 7477 | hCoV-19/England/MILK-991ED4/2020\|EPI_ISL_552085\|20 | ···················· | ···················· | ····················· | ···················· | ···················· | ···················· |
| J | 7478 | hCoV-19/England/MILK-991EE3/2020\|EPI_ISL_552086\|20 | ···················· | ···················· | ····················· | ···················· | ···················· | ···················· |
| J | 7479 | hCoV-19/England/MILK-993230/2020\|EPI_ISL_552087\|20 | ···················· | ···················· | ····················· | ···················· | ···················· | ···················· |
| J | 7480 | hCoV-19/England/MILK-993388/2020\|EPI_ISL_552089\|20 | ···················· | ···················· | ····················· | ···················· | ···················· | ···················· |
| J | 7481 | hCoV-19/England/MILK-993555/2020\|EPI_ISL_552090\|20 | ···················· | ···················· | ····················· | ···················· | ···················· | ···················· |
| J | 7482 | hCoV-19/England/CAMC-991566/2020\|EPI_ISL_552091\|20 | ···················· | ···················· | ····················· | ···················· | ···················· | ···················· |
| J | 7483 | hCoV-19/England/MILK-991EA7/2020\|EPI_ISL_552092\|20 | ···················· | ···················· | ····················· | ···················· | ···················· | ···················· |
| J | 7484 | hCoV-19/England/MILK-99294B/2020\|EPI_ISL_552093\|20 | ···················· | ·A·······A·········· | ····················· | ···················· | ···················· | ···················· |
| J | 7485 | hCoV-19/England/CAMC-991496/2020\|EPI_ISL_552094\|20 | ···················· | ···················· | ····················· | ···················· | ···················· | ···················· |
| J | 7486 | hCoV-19/England/MILK-993106/2020\|EPI_ISL_552096\|20 | ···················· | ···················· | ····················· | ···················· | ···················· | ···················· |
| J | 7487 | hCoV-19/England/MILK-991E3E/2020\|EPI_ISL_552097\|20 | ···················· | ···················· | ····················· | ···················· | ···················· | ···················· |
| J | 7488 | hCoV-19/England/CAMC-9914F0/2020\|EPI_ISL_552098\|20 | ···················· | ···················· | ····················· | ···················· | ···················· | ···················· |
| J | 7489 | hCoV-19/England/MILK-99369E/2020\|EPI_ISL_552099\|20 | ···················· | ···················· | ····················· | ···················· | ···················· | ···················· |
| J | 7490 | hCoV-19/England/ALDP-95A4E4/2020\|EPI_ISL_553358\|20 | ···················· | ···················· | ····················· | ···················· | ···················· | ···················· |
| J | 7491 | hCoV-19/England/ALDP-95D205/2020\|EPI_ISL_553360\|20 | ···················· | ···················· | ····················· | ···················· | ···················· | ···················· |
| J | 7492 | hCoV-19/England/ALDP-955364/2020\|EPI_ISL_553361\|20 | ···················· | ···················· | ····················· | ···················· | ···················· | ···················· |
| J | 7493 | hCoV-19/England/ALDP-954CA6/2020\|EPI_ISL_553362\|20 | ···················· | ···················· | ····················· | ···················· | ···················· | ···················· |
| J | 7494 | hCoV-19/England/ALDP-95A39C/2020\|EPI_ISL_553363\|20 | ···················· | ···················· | ····················· | ···················· | ···················· | ···················· |
| J | 7495 | hCoV-19/England/ALDP-95522B/2020\|EPI_ISL_553364\|20 | ···················· | ···················· | ····················· | ···················· | ···················· | ···················· |
| J | 7496 | hCoV-19/England/ALDP-9550B8/2020\|EPI_ISL_553365\|20 | ···················· | ···················· | ····················· | ···················· | ···················· | ···················· |
| J | 7497 | hCoV-19/England/ALDP-95A4A8/2020\|EPI_ISL_553366\|20 | ···················· | ···················· | ····················· | ···················· | ···················· | ···················· |
| J | 7498 | hCoV-19/England/ALDP-959FB7/2020\|EPI_ISL_553367\|20 | ···················· | ···················· | ····················· | ···················· | ···················· | ···················· |
| J | 7499 | hCoV-19/England/ALDP-95A55A/2020\|EPI_ISL_553368\|20 | ···················· | ···················· | ····················· | ···················· | ···················· | ···················· |
| J | 7500 | hCoV-19/England/ALDP-955601/2020\|EPI_ISL_553369\|20 | ···················· | ···················· | ····················· | ···················· | ···················· | ···················· |
| J | 7501 | hCoV-19/England/ALDP-954D94/2020\|EPI_ISL_553370\|20 | ···················· | ···················· | ····················· | ···················· | ···················· | ···················· |
| J | 7502 | hCoV-19/England/ALDP-95514C/2020\|EPI_ISL_553371\|20 | ···················· | ···················· | ····················· | ···················· | ···················· | ···················· |
| J | 7503 | hCoV-19/England/ALDP-95C87D/2020\|EPI_ISL_553372\|20 | ···················· | ···················· | ····················· | ···················· | ···················· | ···················· |
| J | 7504 | hCoV-19/England/ALDP-9555AA/2020\|EPI_ISL_553373\|20 | ···················· | ···················· | ····················· | ···················· | ···················· | ···················· |
| J | 7505 | hCoV-19/England/ALDP-954A15/2020\|EPI_ISL_553374\|20 | ···················· | ···················· | ····················· | ···················· | ···················· | ···················· |
| J | 7506 | hCoV-19/England/ALDP-95512E/2020\|EPI_ISL_553376\|20 | ···················· | ···················· | ····················· | ···················· | ···················· | ···················· |
| J | 7507 | hCoV-19/England/ALDP-955355/2020\|EPI_ISL_553377\|20 | ···················· | ···················· | ····················· | ···················· | ···················· | ···················· |
| J | 7508 | hCoV-19/England/ALDP-9553A0/2020\|EPI_ISL_553378\|20 | ···················· | ···················· | ····················· | ···················· | ···················· | ···················· |
| J | 7509 | hCoV-19/England/ALDP-95A5B4/2020\|EPI_ISL_553379\|20 | ···················· | ···················· | ····················· | ···················· | ···················· | ···················· |
| J | 7510 | hCoV-19/England/ALDP-94D499/2020\|EPI_ISL_553380\|20 | ···················· | ···················· | ····················· | ···················· | ···················· | ···················· |
| J | 7511 | hCoV-19/England/ALDP-955030/2020\|EPI_ISL_553382\|20 | ···················· | ···················· | ····················· | ···················· | ···················· | ···················· |
| J | 7512 | hCoV-19/England/ALDP-95D1EA/2020\|EPI_ISL_553383\|20 | ···················· | ···················· | ····················· | ···················· | ···················· | ···················· |
| J | 7513 | hCoV-19/England/ALDP-95A3D8/2020\|EPI_ISL_553384\|20 | ···················· | ···················· | ····················· | ···················· | ···················· | ···················· |
| J | 7514 | hCoV-19/England/ALDP-95CF0F/2020\|EPI_ISL_553385\|20 | ···················· | ···················· | ····················· | ···················· | ···················· | ···················· |

| | | | | | | | | |
|---|---|---|---|---|---|---|---|---|
| J | 7515 | hCoV-19/England/ALDP-95D28D/2020\|EPI_ISL_553386\|20 | ···················· | ···················· | ····················· | ···················· | ···················· | ···················· |
| J | 7516 | hCoV-19/England/ALDP-9550D6/2020\|EPI_ISL_553388\|20 | ···················· | ···················· | ····················· | ···················· | ···················· | ···················· |
| J | 7517 | hCoV-19/England/ALDP-95A87F/2020\|EPI_ISL_553390\|20 | ···················· | ···················· | ····················· | ···················· | ···················· | ···················· |
| J | 7518 | hCoV-19/England/ALDP-95505E/2020\|EPI_ISL_553391\|20 | ···················· | ···················· | ····················· | ···················· | ···················· | ···················· |
| J | 7519 | hCoV-19/England/ALDP-95A53C/2020\|EPI_ISL_553393\|20 | ···················· | ···················· | ····················· | ···················· | ···················· | ···················· |
| J | 7520 | hCoV-19/England/ALDP-95A806/2020\|EPI_ISL_553394\|20 | ···················· | ···················· | ····················· | ···················· | ···················· | ···················· |
| J | 7521 | hCoV-19/England/ALDP-954C2E/2020\|EPI_ISL_553395\|20 | ···················· | ···················· | ····················· | ···················· | ···················· | ···················· |
| J | 7522 | hCoV-19/England/ALDP-95C93E/2020\|EPI_ISL_553396\|20 | ···················· | ···················· | ····················· | ···················· | ···················· | ···················· |
| J | 7523 | hCoV-19/England/ALDP-9549BE/2020\|EPI_ISL_553397\|20 | ···················· | ···················· | ····················· | ···················· | ···················· | ···················· |
| J | 7524 | hCoV-19/England/ALDP-95A50F/2020\|EPI_ISL_553399\|20 | ···················· | ···················· | ····················· | ···················· | ···················· | ···················· |
| J | 7525 | hCoV-19/England/ALDP-9557E0/2020\|EPI_ISL_553400\|20 | ···················· | ···················· | ····················· | ···················· | ···················· | ···················· |
| J | 7526 | hCoV-19/England/ALDP-954B21/2020\|EPI_ISL_553405\|20 | ···················· | ·A·················· | ····················· | ···················· | ···················· | ···················· |
| J | 7527 | hCoV-19/England/ALDP-9549FA/2020\|EPI_ISL_553406\|20 | ···················· | ···················· | ····················· | ···················· | ···················· | ···················· |
| J | 7528 | hCoV-19/England/ALDP-95556E/2020\|EPI_ISL_553408\|20 | ···················· | ···················· | ····················· | ···················· | ···················· | ···················· |
| J | 7529 | hCoV-19/England/ALDP-955540/2020\|EPI_ISL_553409\|20 | ···················· | ···················· | ····················· | ···················· | ···················· | ···················· |
| J | 7530 | hCoV-19/England/ALDP-95CEF3/2020\|EPI_ISL_553410\|20 | ···················· | ···················· | ····················· | ···················· | ···················· | ···················· |
| J | 7531 | hCoV-19/England/ALDP-954B9A/2020\|EPI_ISL_553411\|20 | ···················· | ···················· | ····················· | ···················· | ···················· | ···················· |
| J | 7532 | hCoV-19/England/ALDP-955337/2020\|EPI_ISL_553500\|20 | ···················· | ···················· | ····················· | ···················· | ···················· | ···················· |
| J | 7533 | hCoV-19/England/ALDP-954C6A/2020\|EPI_ISL_553501\|20 | ···················· | ···················· | ····················· | ···················· | ···················· | ···················· |
| J | 7534 | hCoV-19/England/ALDP-954DEF/2020\|EPI_ISL_553502\|20 | ···················· | ···················· | ····················· | ···················· | ···················· | ···················· |
| J | 7535 | hCoV-19/England/ALDP-954BA9/2020\|EPI_ISL_553503\|20 | ···················· | ···················· | ····················· | ···················· | ···················· | ···················· |
| J | 7536 | hCoV-19/England/ALDP-95CF4B/2020\|EPI_ISL_553504\|20 | ···················· | ···················· | ····················· | ···················· | ···················· | ···················· |
| J | 7537 | hCoV-19/England/ALDP-94D47B/2020\|EPI_ISL_553505\|20 | ···················· | ···················· | ····················· | ···················· | ···················· | ···················· |
| J | 7538 | hCoV-19/England/ALDP-95A51E/2020\|EPI_ISL_553506\|20 | ···················· | ···················· | ····················· | ···················· | ···················· | ···················· |
| J | 7539 | hCoV-19/England/ALDP-959FE4/2020\|EPI_ISL_553507\|20 | ···················· | ···················· | ····················· | ···················· | ···················· | ···················· |
| J | 7540 | hCoV-19/England/ALDP-944526/2020\|EPI_ISL_554688\|20 | ···················· | ·A·················· | ····················· | ···················· | ···················· | ···················· |
| J | 7541 | hCoV-19/England/ALDP-944474/2020\|EPI_ISL_554689\|20 | ···················· | ···················· | ····················· | ···················· | ···················· | ···················· |
| J | 7542 | hCoV-19/England/MILK-9589BA/2020\|EPI_ISL_554690\|20 | ···················· | ···················· | ····················· | ···················· | ···················· | ···················· |
| J | 7543 | hCoV-19/England/MILK-958510/2020\|EPI_ISL_554691\|20 | ···················· | ···················· | ····················· | ···················· | ···················· | ···················· |
| J | 7544 | hCoV-19/England/ALDP-9441F5/2020\|EPI_ISL_554692\|20 | ···················· | ···················· | ····················· | ···················· | ···················· | ···················· |
| J | 7545 | hCoV-19/England/ALDP-944052/2020\|EPI_ISL_554694\|20 | ···················· | ···················· | ····················· | ···················· | ···················· | ···················· |
| J | 7546 | hCoV-19/England/MILK-95846E/2020\|EPI_ISL_554695\|20 | ···················· | ···················· | ····················· | ···················· | ···················· | ···················· |
| J | 7547 | hCoV-19/England/ALDP-944623/2020\|EPI_ISL_554696\|20 | ···················· | ···················· | ····················· | ···················· | ···················· | ···················· |
| J | 7548 | hCoV-19/England/ALDP-94432C/2020\|EPI_ISL_554697\|20 | ···················· | ···················· | ····················· | ···················· | ···················· | ···················· |
| J | 7549 | hCoV-19/England/MILK-958B3C/2020\|EPI_ISL_554698\|20 | ···················· | ···················· | ····················· | ···················· | ···················· | ···················· |
| J | 7550 | hCoV-19/England/MILK-958422/2020\|EPI_ISL_554721\|20 | ···················· | ···················· | ····················· | ···················· | ···················· | ···················· |
| J | 7551 | hCoV-19/England/MILK-958941/2020\|EPI_ISL_554722\|20 | ···················· | ···················· | ····················· | ···················· | ···················· | ···················· |
| J | 7552 | hCoV-19/England/MILK-958088/2020\|EPI_ISL_554723\|20 | ···················· | ···················· | ····················· | ···················· | ···················· | ···················· |
| J | 7553 | hCoV-19/England/MILK-957D64/2020\|EPI_ISL_554724\|20 | ···················· | ···················· | ····················· | ···················· | ···················· | ···················· |
| J | 7554 | hCoV-19/England/ALDP-9440CB/2020\|EPI_ISL_554725\|20 | ···················· | ···················· | ····················· | ···················· | ···················· | ···················· |
| J | 7555 | hCoV-19/England/ALDP-94466F/2020\|EPI_ISL_554726\|20 | ···················· | ···················· | ····················· | ···················· | ···················· | ···················· |
| J | 7556 | hCoV-19/England/ALDP-944298/2020\|EPI_ISL_554727\|20 | ···················· | ···················· | ····················· | ···················· | ···················· | ···················· |
| J | 7557 | hCoV-19/England/MILK-958844/2020\|EPI_ISL_554729\|20 | ···················· | ···················· | ····················· | ···················· | ···················· | ···················· |
| J | 7558 | hCoV-19/England/ALDP-944483/2020\|EPI_ISL_554730\|20 | ···················· | ···················· | ····················· | ···················· | ···················· | ···················· |
| J | 7559 | hCoV-19/England/MILK-95862C/2020\|EPI_ISL_554731\|20 | ···················· | ···················· | ····················· | ···················· | ···················· | ···················· |
| J | 7560 | hCoV-19/Wales/MILK-958659/2020\|EPI_ISL_554734\|2020 | ···················· | ···················· | ····················· | ···················· | ···················· | ···················· |
| J | 7561 | hCoV-19/England/MILK-958149/2020\|EPI_ISL_554737\|20 | ···················· | ···················· | ····················· | ···················· | ···················· | ···················· |
| J | 7562 | hCoV-19/England/MILK-9580E2/2020\|EPI_ISL_554738\|20 | ···················· | ···················· | ····················· | ···················· | ···················· | ···················· |
| J | 7563 | hCoV-19/England/MILK-9589F6/2020\|EPI_ISL_554739\|20 | ···················· | ···················· | ····················· | ···················· | ···················· | ···················· |
| J | 7564 | hCoV-19/England/MILK-959940/2020\|EPI_ISL_554740\|20 | ···················· | ···················· | ····················· | ···················· | ···················· | ···················· |
| J | 7565 | hCoV-19/England/ALDP-943F92/2020\|EPI_ISL_554741\|20 | ···················· | ···················· | ····················· | ···················· | ···················· | ···················· |
| J | 7566 | hCoV-19/England/MILK-9586FF/2020\|EPI_ISL_554831\|20 | ···················· | ·A·················· | ····················· | ···················· | ···················· | ···················· |
| J | 7567 | hCoV-19/England/ALDP-94440B/2020\|EPI_ISL_554832\|20 | ···················· | ·A·················· | ····················· | ···················· | ···················· | ···················· |
| J | 7568 | hCoV-19/England/ALDP-944429/2020\|EPI_ISL_554833\|20 | ···················· | ···················· | ····················· | ···················· | ···················· | ···················· |
| J | 7569 | hCoV-19/England/ALDP-944043/2020\|EPI_ISL_554834\|20 | ···················· | ···················· | ····················· | ···················· | ···················· | ···················· |
| J | 7570 | hCoV-19/England/MILK-958501/2020\|EPI_ISL_554835\|20 | ···················· | ···················· | ····················· | ···················· | ···················· | ···················· |
| J | 7571 | hCoV-19/England/ALDP-944025/2020\|EPI_ISL_554836\|20 | ···················· | ···················· | ····················· | ···················· | ···················· | ···················· |
| J | 7572 | hCoV-19/England/ALDP-943FB0/2020\|EPI_ISL_554837\|20 | ···················· | ···················· | ····················· | ···················· | ···················· | ···················· |
| J | 7573 | hCoV-19/England/MILK-95802E/2020\|EPI_ISL_554838\|20 | ···················· | ···················· | ····················· | ···················· | ···················· | ···················· |
| J | 7574 | hCoV-19/England/ALDP-68E0E6/2020\|EPI_ISL_558200\|20 | ···················· | ···················· | ····················· | ···················· | ···················· | ···················· |
| J | 7575 | hCoV-19/England/ALDP-692780/2020\|EPI_ISL_558201\|20 | ···················· | ···················· | ····················· | ···················· | ···················· | ···················· |
| J | 7576 | hCoV-19/England/ALDP-690193/2020\|EPI_ISL_558202\|20 | ···················· | ···················· | ····················· | ···················· | ···················· | ···················· |
| J | 7577 | hCoV-19/England/ALDP-647D3D/2020\|EPI_ISL_558203\|20 | ···················· | ···················· | ····················· | ···················· | ···················· | ···················· |
| J | 7578 | hCoV-19/England/ALDP-66A9D6/2020\|EPI_ISL_558205\|20 | ···················· | ···················· | ····················· | ···················· | ···················· | ···················· |
| J | 7579 | hCoV-19/England/ALDP-68F13D/2020\|EPI_ISL_558206\|20 | ···················· | ···················· | ····················· | ···················· | ···················· | ···················· |
| J | 7580 | hCoV-19/England/ALDP-647F46/2020\|EPI_ISL_558207\|20 | ···················· | ···················· | ····················· | ···················· | ···················· | ···················· |
| J | 7581 | hCoV-19/England/ALDP-691C19/2020\|EPI_ISL_558209\|20 | ···················· | ···················· | ····················· | ···················· | ···················· | ···················· |
| J | 7582 | hCoV-19/England/ALDP-67654C/2020\|EPI_ISL_558210\|20 | ···················· | ···················· | ····················· | ···················· | ···················· | ···················· |
| J | 7583 | hCoV-19/England/ALDP-652045/2020\|EPI_ISL_558212\|20 | ···················· | ······A············· | ····················· | ···················· | ···················· | ···················· |
| J | 7584 | hCoV-19/England/ALDP-64E0F1/2020\|EPI_ISL_558213\|20 | ···················· | ···················· | ····················· | ···················· | ···················· | ···················· |
| J | 7585 | hCoV-19/England/ALDP-68A484/2020\|EPI_ISL_558216\|20 | ···················· | ···················· | ····················· | ···················· | ···················· | ···················· |
| J | 7586 | hCoV-19/England/ALDP-693673/2020\|EPI_ISL_558217\|20 | ···················· | ···················· | ····················· | ···················· | ···················· | ···················· |
| J | 7587 | hCoV-19/England/ALDP-658944/2020\|EPI_ISL_558219\|20 | ···················· | ···················· | ····················· | ···················· | ···················· | ···················· |
| J | 7588 | hCoV-19/England/ALDP-66B638/2020\|EPI_ISL_558220\|20 | ···················· | ···················· | ····················· | ···················· | ···················· | ···················· |
| J | 7589 | hCoV-19/England/ALDP-66B1BF/2020\|EPI_ISL_558221\|20 | ···················· | ···················· | ····················· | ···················· | ···················· | ···················· |
| J | 7590 | hCoV-19/England/ALDP-673B98/2020\|EPI_ISL_558223\|20 | ···················· | ···················· | ····················· | ···················· | ···················· | ···················· |
| J | 7591 | hCoV-19/England/ALDP-66F115/2020\|EPI_ISL_558226\|20 | ···················· | ···················· | ····················· | ···················· | ···················· | ···················· |
| J | 7592 | hCoV-19/England/ALDP-68C3FE/2020\|EPI_ISL_558227\|20 | ···················· | ···················· | ····················· | ···················· | ···················· | ···················· |
| J | 7593 | hCoV-19/England/ALDP-653017/2020\|EPI_ISL_558228\|20 | ···················· | ···················· | ····················· | ···················· | ···················· | ···················· |
| J | 7594 | hCoV-19/England/ALDP-691499/2020\|EPI_ISL_558230\|20 | ···················· | ···················· | ····················· | ···················· | ···················· | ···················· |

J 7595 hCoV-19/England/ALDP-68E0C8/2020|EPI_ISL_558231|20 ···················· ···················· ····················· ···················· ···················· ····················
J 7596 hCoV-19/England/ALDP-66F379/2020|EPI_ISL_558232|20 ···················· ···················· ····················· ···················· ···················· ····················
J 7597 hCoV-19/England/ALDP-66C822/2020|EPI_ISL_558233|20 ···················· ···················· ····················· ···················· ···················· ····················
J 7598 hCoV-19/England/MILK-66F889/2020|EPI_ISL_558234|20 ···················· ···················· ····················· ···················· ···················· ····················
J 7599 hCoV-19/England/ALDP-64831C/2020|EPI_ISL_558235|20 ···················· ···················· ····················· ···················· ···················· ····················
J 7600 hCoV-19/England/ALDP-68EFBC/2020|EPI_ISL_558236|20 ···················· ···················· ····················· ···················· ···················· ····················
J 7601 hCoV-19/England/ALDP-59AC8A/2020|EPI_ISL_558238|20 ···················· ···················· ····················· ···················· ···················· ····················
J 7602 hCoV-19/England/MILK-5642D2/2020|EPI_ISL_558239|20 ···················· ···················· ····················· ···················· ···················· ····················
J 7603 hCoV-19/England/ALDP-59AC20/2020|EPI_ISL_558240|20 ···················· ···················· ····················· ···················· ···················· ····················
J 7604 hCoV-19/England/MILK-597072/2020|EPI_ISL_558241|20 ···················· ···················· ···········T········· ···················· ···················· ····················
J 7605 hCoV-19/Wales/MILK-55D47C/2020|EPI_ISL_558242|2020 ···················· ···················· ····················· ···················· ···················· ····················
J 7606 hCoV-19/England/MILK-559ABA/2020|EPI_ISL_558243|20 ···················· ···················· ····················· ···················· ···················· ····················
J 7607 hCoV-19/England/ALDP-59BB9B/2020|EPI_ISL_558244|20 ···················· ···················· ····················· ···················· ···················· ····················
J 7608 hCoV-19/England/MILK-559DA2/2020|EPI_ISL_558245|20 ···················· ···················· ····················· ···················· ···················· ····················
J 7609 hCoV-19/England/MILK-577FA8/2020|EPI_ISL_558246|20 ···················· ···················· ····················· ···················· ···················· ····················
J 7610 hCoV-19/England/MILK-57B0FC/2020|EPI_ISL_558247|20 ···················· ···················· ····················· ···················· ···················· ····················
J 7611 hCoV-19/England/ALDP-4A8E57/2020|EPI_ISL_559500|20 ···················· ···················· ····················· ···················· ···················· ····················
J 7612 hCoV-19/England/ALDP-49CB04/2020|EPI_ISL_559501|20 ···················· ···················· ····················· ···················· ···················· ····················
J 7613 hCoV-19/England/ALDP-4AE1B2/2020|EPI_ISL_559502|20 ···················· ···················· ····················· ···················· ···················· ····················
J 7614 hCoV-19/England/ALDP-4A44A4/2020|EPI_ISL_559505|20 ···················· ···················· ····················· ···················· ···················· ····················
J 7615 hCoV-19/England/ALDP-4A80BB/2020|EPI_ISL_559506|20 ···················· ···················· ····················· ···················· ···················· ····················
J 7616 hCoV-19/England/ALDP-4A69D0/2020|EPI_ISL_559507|20 ···················· ···················· ····················· ···················· ···················· ····················
J 7617 hCoV-19/England/ALDP-4ACD0B/2020|EPI_ISL_559508|20 ···················· ···················· ····················· ···················· ···················· ····················
J 7618 hCoV-19/England/ALDP-4A5B53/2020|EPI_ISL_559509|20 ···················· ···················· ····················· ···················· ···················· ····················
J 7619 hCoV-19/England/ALDP-4A7A09/2020|EPI_ISL_559510|20 ···················· ···················· ····················· ···················· ···················· ····················
J 7620 hCoV-19/England/ALDP-4AC776/2020|EPI_ISL_559511|20 ···················· ···················· ····················· ···················· ···················· ····················
J 7621 hCoV-19/England/ALDP-4AE0C4/2020|EPI_ISL_559512|20 ···················· ···················· ····················· ···················· ···················· ····················
J 7622 hCoV-19/England/ALDP-4A922D/2020|EPI_ISL_559513|20 ···················· ···················· ····················· ···················· ···················· ····················
J 7623 hCoV-19/England/ALDP-4AD0E3/2020|EPI_ISL_559514|20 ···················· ···················· ····················· ···················· ···················· ····················
J 7624 hCoV-19/England/ALDP-4A9339/2020|EPI_ISL_559515|20 ···················· ···················· ····················· ···················· ···················· ····················
J 7625 hCoV-19/England/ALDP-4A5EB4/2020|EPI_ISL_559516|20 ···················· ···················· ····················· ···················· ···················· ····················
J 7626 hCoV-19/England/ALDP-4A37CA/2020|EPI_ISL_559518|20 ···················· ···················· ····················· ···················· ···················· ····················
J 7627 hCoV-19/England/ALDP-49CE38/2020|EPI_ISL_559519|20 ···················· ···················· ····················· ···················· ···················· ····················
J 7628 hCoV-19/England/ALDP-4A48D5/2020|EPI_ISL_559520|20 ···················· ···················· ····················· ···················· ···················· ····················
J 7629 hCoV-19/England/ALDP-4A628A/2020|EPI_ISL_559521|20 ···················· ···················· ····················· ···················· ···················· ····················
J 7630 hCoV-19/England/ALDP-4A5E4B/2020|EPI_ISL_559523|20 ···················· ···················· ····················· ···················· ···················· ····················
J 7631 hCoV-19/England/ALDP-4AA16B/2020|EPI_ISL_559525|20 ···················· ···················· ····················· ···················· ···················· ····················
J 7632 hCoV-19/England/ALDP-49E355/2020|EPI_ISL_559526|20 ···················· ···················· ····················· ···················· ···················· ····················
J 7633 hCoV-19/England/ALDP-4AE0D3/2020|EPI_ISL_559527|20 ···················· ···················· ····················· ···················· ···················· ····················
J 7634 hCoV-19/England/ALDP-4A9885/2020|EPI_ISL_559528|20 ···················· ···················· ····················· ···················· ···················· ····················
J 7635 hCoV-19/England/ALDP-4AC20C/2020|EPI_ISL_559529|20 ···················· ···················· ····················· ···················· ···················· ····················
J 7636 hCoV-19/England/ALDP-4A44E0/2020|EPI_ISL_559530|20 ···················· ···················· ····················· ···················· ···················· ····················
J 7637 hCoV-19/England/ALDP-49C16C/2020|EPI_ISL_559531|20 ···················· ···················· ····················· ···················· ···················· ····················
J 7638 hCoV-19/England/ALDP-4A52E5/2020|EPI_ISL_559532|20 ···················· ···················· ····················· ···················· ···················· ····················
J 7639 hCoV-19/England/ALDP-4A8A44/2020|EPI_ISL_559533|20 ···················· ···················· ····················· ···················· ···················· ····················
J 7640 hCoV-19/England/ALDP-4A7016/2020|EPI_ISL_559534|20 ···················· ···················· ····················· ···················· ···················· ····················
J 7641 hCoV-19/England/ALDP-4A8376/2020|EPI_ISL_559535|20 ···················· ···················· ····················· ···················· ···················· ····················
J 7642 hCoV-19/England/ALDP-4A5607/2020|EPI_ISL_559536|20 ···················· ···················· ····················· ···················· ···················· ····················
J 7643 hCoV-19/England/ALDP-4A4C9D/2020|EPI_ISL_559537|20 ···················· ···················· ····················· ···················· ···················· ····················
J 7644 hCoV-19/England/ALDP-49EDDF/2020|EPI_ISL_559538|20 ···················· ···················· ····················· ···················· ···················· ····················
J 7645 hCoV-19/England/ALDP-4ACDA1/2020|EPI_ISL_559539|20 ···················· ···················· ····················· ···················· ···················· ····················
J 7646 hCoV-19/England/ALDP-4A76E7/2020|EPI_ISL_559540|20 ···················· ···················· ····················· ···················· ···················· ····················
J 7647 hCoV-19/England/ALDP-49E980/2020|EPI_ISL_559541|20 ···················· ···················· ····················· ···················· ···················· ····················
J 7648 hCoV-19/England/ALDP-4A8570/2020|EPI_ISL_559542|20 ···················· ···················· ····················· ···················· ···················· ····················
J 7649 hCoV-19/England/ALDP-4AE352/2020|EPI_ISL_559543|20 ···················· ···················· ····················· ···················· ···················· ····················
J 7650 hCoV-19/England/ALDP-4A7456/2020|EPI_ISL_559546|20 ···················· ···················· ····················· ···················· ···················· ····················
J 7651 hCoV-19/England/ALDP-49CD2C/2020|EPI_ISL_559547|20 ···················· ···················· ····················· ···················· ···················· ····················
J 7652 hCoV-19/England/ALDP-4AA5E7/2020|EPI_ISL_559548|20 ···················· ···················· ····················· ···················· ···················· ····················
J 7653 hCoV-19/England/ALDP-497F2B/2020|EPI_ISL_559550|20 ···················· ···················· ····················· ···················· ···················· ····················
J 7654 hCoV-19/England/ALDP-4AC1E1/2020|EPI_ISL_559551|20 ···················· ···················· ····················· ···················· ···················· ····················
J 7655 hCoV-19/England/ALDP-4A6008/2020|EPI_ISL_559552|20 ···················· ···················· ····················· ···················· ···················· ····················
J 7656 hCoV-19/England/ALDP-4A975B/2020|EPI_ISL_559553|20 ···················· ···················· ····················· ···················· ···················· ····················
J 7657 hCoV-19/England/ALDP-4AC925/2020|EPI_ISL_559554|20 ···················· ···················· ····················· ···················· ···················· ····················
J 7658 hCoV-19/England/ALDP-4A5A92/2020|EPI_ISL_559555|20 ···················· ···················· ····················· ···················· ···················· ····················
J 7659 hCoV-19/England/ALDP-4A5C14/2020|EPI_ISL_559556|20 ···················· ···················· ····················· ···················· ···················· ····················
J 7660 hCoV-19/England/ALDP-4ACC95/2020|EPI_ISL_559557|20 ···················· ···················· ····················· ···················· ···················· ····················
J 7661 hCoV-19/England/ALDP-49DEBF/2020|EPI_ISL_559558|20 ···················· ···················· ····················· ···················· ···················· ····················
J 7662 hCoV-19/England/ALDP-4ADE8F/2020|EPI_ISL_559559|20 ···················· ···················· ····················· ···················· ···················· ····················
J 7663 hCoV-19/England/ALDP-4A52C7/2020|EPI_ISL_559560|20 ···················· ···················· ····················· ···················· ···················· ····················
J 7664 hCoV-19/England/ALDP-49CD0E/2020|EPI_ISL_559561|20 ···················· ···················· ····················· ···················· ···················· ····················
J 7665 hCoV-19/England/ALDP-4A437A/2020|EPI_ISL_559562|20 ···················· ···················· ····················· ···················· ···················· ····················
J 7666 hCoV-19/England/ALDP-4A4213/2020|EPI_ISL_559563|20 ···················· ···················· ····················· ···················· ···················· ····················
J 7667 hCoV-19/England/ALDP-4A7948/2020|EPI_ISL_559564|20 ···················· ···················· ····················· ···················· ···················· ····················
J 7668 hCoV-19/England/ALDP-49D23B/2020|EPI_ISL_559565|20 ···················· ···················· ····················· ···················· ···················· ····················
J 7669 hCoV-19/England/ALDP-49F0B7/2020|EPI_ISL_559566|20 ···················· ···················· ····················· ···················· ···················· ····················
J 7670 hCoV-19/England/ALDP-4A9DC2/2020|EPI_ISL_559567|20 ···················· ···················· ····················· ···················· ···················· ····················
J 7671 hCoV-19/England/ALDP-4A519D/2020|EPI_ISL_559568|20 ···················· ···················· ····················· ···················· ···················· ····················
J 7672 hCoV-19/England/ALDP-4AD863/2020|EPI_ISL_559569|20 ···················· ···················· ····················· ···················· ···················· ····················
J 7673 hCoV-19/England/ALDP-49D24A/2020|EPI_ISL_559570|20 ···················· ···················· ····················· ···················· ···················· ····················
J 7674 hCoV-19/England/ALDP-4AAE28/2020|EPI_ISL_559571|20 ···················· ·A·················· ····················· ···················· ···················· ····················

| | | | | | | | | |
|---|---|---|---|---|---|---|---|---|
| J | 7675 | hCoV-19/England/ALDP-4AA81B/2020\|EPI_ISL_559572\|20 | ···················· | ···················· | ····················· | ···················· | ···················· | ···················· |
| J | 7676 | hCoV-19/England/ALDP-4A35C0/2020\|EPI_ISL_559573\|20 | ···················· | ···················· | ····················· | ···················· | ···················· | ···················· |
| J | 7677 | hCoV-19/England/ALDP-4AA927/2020\|EPI_ISL_559574\|20 | ···················· | ···················· | ····················· | ···················· | ···················· | ···················· |
| J | 7678 | hCoV-19/England/ALDP-4AE06A/2020\|EPI_ISL_559577\|20 | ···················· | ···················· | ····················· | ···················· | ···················· | ···················· |
| J | 7679 | hCoV-19/England/ALDP-4A766F/2020\|EPI_ISL_559578\|20 | ···················· | ···················· | ····················· | ···················· | ···················· | ···················· |
| J | 7680 | hCoV-19/England/ALDP-4AE835/2020\|EPI_ISL_559579\|20 | ···················· | ···················· | ····················· | ···················· | ···················· | ···················· |
| J | 7681 | hCoV-19/England/MILK-992EB5/2020\|EPI_ISL_552100\|20 | ···················· | ···················· | ····················· | ···················· | ······T············· | ···················· |
| J | 7682 | hCoV-19/England/MILK-991EB6/2020\|EPI_ISL_552101\|20 | ···················· | ···················· | ····················· | ···················· | ···················· | ···················· |
| J | 7683 | hCoV-19/England/MILK-992A0C/2020\|EPI_ISL_552102\|20 | ···················· | ···················· | ····················· | ···················· | ···················· | ···················· |
| J | 7684 | hCoV-19/England/MILK-993494/2020\|EPI_ISL_552103\|20 | ···················· | ···················· | ····················· | ···················· | ···················· | ···················· |
| J | 7685 | hCoV-19/England/MILK-992FB2/2020\|EPI_ISL_552104\|20 | ···················· | ·A·················· | ····················· | ···················· | ···················· | ···················· |
| J | 7686 | hCoV-19/England/MILK-993722/2020\|EPI_ISL_552107\|20 | ···················· | ···················· | ····················· | ···················· | ···················· | ···················· |
| J | 7687 | hCoV-19/England/MILK-992B45/2020\|EPI_ISL_552108\|20 | ···················· | ···················· | ····················· | ···················· | ···················· | ···················· |
| J | 7688 | hCoV-19/England/MILK-9930FA/2020\|EPI_ISL_552109\|20 | ···················· | ···················· | ····················· | ···················· | ···················· | ···················· |
| J | 7689 | hCoV-19/England/MILK-9926DB/2020\|EPI_ISL_552110\|20 | ···················· | ···················· | ····················· | ···················· | ···················· | ···················· |
| J | 7690 | hCoV-19/England/MILK-9936E9/2020\|EPI_ISL_552111\|20 | ···················· | ···················· | ····················· | ···················· | ···················· | ···················· |
| J | 7691 | hCoV-19/England/MILK-993582/2020\|EPI_ISL_552112\|20 | ···················· | ···················· | ····················· | ···················· | ···················· | ···················· |
| J | 7692 | hCoV-19/England/MILK-99279C/2020\|EPI_ISL_552113\|20 | ···················· | ···················· | ····················· | ···················· | ···················· | ···················· |
| J | 7693 | hCoV-19/England/MILK-993625/2020\|EPI_ISL_552114\|20 | ···················· | ···················· | ····················· | ···················· | ···················· | ···················· |
| J | 7694 | hCoV-19/England/MILK-992987/2020\|EPI_ISL_552115\|20 | ···················· | ·A·······A·········· | ····················· | ···················· | ···················· | ···················· |
| J | 7695 | hCoV-19/England/MILK-993564/2020\|EPI_ISL_552116\|20 | ···················· | ···················· | ····················· | ···················· | ···················· | ···················· |
| J | 7696 | hCoV-19/Wales/MILK-99368F/2020\|EPI_ISL_552117\|2020 | ···················· | ···················· | ····················· | ···················· | ···················· | ···················· |
| J | 7697 | hCoV-19/England/MILK-99333D/2020\|EPI_ISL_552118\|20 | ···················· | ···················· | ····················· | ···················· | ···················· | ···················· |
| J | 7698 | hCoV-19/England/MILK-993528/2020\|EPI_ISL_552119\|20 | ···················· | ···················· | ····················· | ···················· | ···················· | ···················· |
| J | 7699 | hCoV-19/England/MILK-992B90/2020\|EPI_ISL_552120\|20 | ···················· | ·A·················· | ····················· | ···················· | ···················· | ···A················ |
| J | 7700 | hCoV-19/England/MILK-99319D/2020\|EPI_ISL_552121\|20 | ···················· | ···················· | ····················· | ···················· | ···················· | ···················· |
| J | 7701 | hCoV-19/England/MILK-99288A/2020\|EPI_ISL_552122\|20 | ···················· | ···················· | ····················· | ···················· | ···················· | ···················· |
| J | 7702 | hCoV-19/England/MILK-992D21/2020\|EPI_ISL_552123\|20 | ···················· | ···················· | ····················· | ···················· | ···················· | ···················· |
| J | 7703 | hCoV-19/England/MILK-992E88/2020\|EPI_ISL_552127\|20 | ···················· | ·A·················· | ····················· | ···················· | ···················· | ···················· |
| J | 7704 | hCoV-19/England/MILK-991E6B/2020\|EPI_ISL_552128\|20 | ···················· | ···················· | ····················· | ···················· | ···················· | ···················· |
| J | 7705 | hCoV-19/England/MILK-992899/2020\|EPI_ISL_552129\|20 | ···················· | ···················· | ····················· | ···················· | ···················· | ···················· |
| J | 7706 | hCoV-19/England/MILK-992C7F/2020\|EPI_ISL_552130\|20 | ···················· | ···················· | ····················· | ···················· | ······T············· | ···················· |
| J | 7707 | hCoV-19/England/MILK-992617/2020\|EPI_ISL_552131\|20 | ···················· | ···················· | ····················· | ···················· | ···················· | ···················· |
| J | 7708 | hCoV-19/Wales/MILK-9931CA/2020\|EPI_ISL_552132\|2020 | ···················· | ···················· | ····················· | ···················· | ···················· | ···················· |
| J | 7709 | hCoV-19/England/MILK-993643/2020\|EPI_ISL_552133\|20 | ···················· | ···················· | ····················· | ···················· | ···················· | ···················· |
| J | 7710 | hCoV-19/England/MILK-993212/2020\|EPI_ISL_552134\|20 | ···················· | ···················· | ····················· | ···················· | ···················· | ···················· |
| J | 7711 | hCoV-19/England/MILK-99324F/2020\|EPI_ISL_552135\|20 | ···················· | ···················· | ····················· | ···················· | ···················· | ···················· |
| J | 7712 | hCoV-19/England/MILK-99285D/2020\|EPI_ISL_552136\|20 | ···················· | ···················· | ····················· | ···················· | ···················· | ···················· |
| J | 7713 | hCoV-19/England/MILK-992EC4/2020\|EPI_ISL_552137\|20 | ···················· | ·A·················· | ····················· | ···················· | ···················· | ···A················ |
| J | 7714 | hCoV-19/England/MILK-993652/2020\|EPI_ISL_552138\|20 | ···················· | ···················· | ····················· | ···················· | ···················· | ···················· |
| J | 7715 | hCoV-19/England/MILK-9933E2/2020\|EPI_ISL_552139\|20 | ···················· | ···················· | ····················· | ···················· | ···················· | ···················· |
| J | 7716 | hCoV-19/England/MILK-99350A/2020\|EPI_ISL_552140\|20 | ···················· | ···················· | ····················· | ···················· | ···················· | ···················· |
| J | 7717 | hCoV-19/England/MILK-992E6A/2020\|EPI_ISL_552141\|20 | ···················· | ···················· | ····················· | ···················· | ···················· | ···················· |
| J | 7718 | hCoV-19/England/MILK-992A84/2020\|EPI_ISL_552142\|20 | ···················· | ···················· | ····················· | ···················· | ···················· | ···················· |
| J | 7719 | hCoV-19/England/MILK-993485/2020\|EPI_ISL_552143\|20 | ···················· | ···················· | ····················· | ···················· | ···················· | ···················· |
| J | 7720 | hCoV-19/Wales/MILK-9931BB/2020\|EPI_ISL_552144\|2020 | ···················· | ···················· | ····················· | ···················· | ···················· | ···················· |
| J | 7721 | hCoV-19/Wales/MILK-992705/2020\|EPI_ISL_552145\|2020 | ···················· | ···················· | ····················· | ···················· | ···················· | ···················· |
| J | 7722 | hCoV-19/England/MILK-992D5E/2020\|EPI_ISL_552146\|20 | ···················· | ···················· | ····················· | ···················· | ···················· | ···················· |
| J | 7723 | hCoV-19/England/ALDP-955285/2020\|EPI_ISL_553401\|20 | ···················· | ···················· | ····················· | ···················· | ···················· | ···················· |
| J | 7724 | hCoV-19/England/ALDP-954C3D/2020\|EPI_ISL_553402\|20 | ···················· | ···················· | ····················· | ···················· | ···················· | ···················· |
| J | 7725 | hCoV-19/England/ALDP-95A578/2020\|EPI_ISL_553403\|20 | ···················· | ···················· | ····················· | ···················· | ···················· | ···················· |
| J | 7726 | hCoV-19/England/ALDP-954C79/2020\|EPI_ISL_553404\|20 | ···················· | ···················· | ····················· | ···················· | ···················· | ···················· |
| J | 7727 | hCoV-19/England/ALDP-95507C/2020\|EPI_ISL_553412\|20 | ···················· | ···················· | ····················· | ···················· | ···················· | ···················· |
| J | 7728 | hCoV-19/England/ALDP-954D85/2020\|EPI_ISL_553413\|20 | ···················· | ···················· | ····················· | ···················· | ···················· | ···················· |
| J | 7729 | hCoV-19/England/ALDP-95C8F5/2020\|EPI_ISL_553414\|20 | ···················· | ···················· | ····················· | ···················· | ···················· | ···················· |
| J | 7730 | hCoV-19/England/ALDP-95A4F3/2020\|EPI_ISL_553415\|20 | ···················· | ···················· | ····················· | ···················· | ···················· | ···················· |
| J | 7731 | hCoV-19/England/ALDP-954FBC/2020\|EPI_ISL_553416\|20 | ···················· | ···················· | ····················· | ···················· | ···················· | ···················· |
| J | 7732 | hCoV-19/England/ALDP-95A4C6/2020\|EPI_ISL_553417\|20 | ···················· | ···················· | ····················· | ···················· | ···················· | ···················· |
| J | 7733 | hCoV-19/England/ALDP-954AD9/2020\|EPI_ISL_553418\|20 | ···················· | ···················· | ····················· | ···················· | ···················· | ···················· |
| J | 7734 | hCoV-19/England/ALDP-95C9A7/2020\|EPI_ISL_553419\|20 | ···················· | ···················· | ····················· | ···················· | ···················· | ···················· |
| J | 7735 | hCoV-19/England/ALDP-954DB2/2020\|EPI_ISL_553422\|20 | ···················· | ···················· | ····················· | ···················· | ···················· | ···················· |
| J | 7736 | hCoV-19/England/ALDP-954D58/2020\|EPI_ISL_553423\|20 | ···················· | ···················· | ····················· | ···················· | ···················· | ···················· |
| J | 7737 | hCoV-19/England/ALDP-95A54B/2020\|EPI_ISL_553424\|20 | ···················· | ···················· | ····················· | ···················· | ···················· | ···················· |
| J | 7738 | hCoV-19/England/ALDP-954EA0/2020\|EPI_ISL_553425\|20 | ···················· | ···················· | ····················· | ···················· | ···················· | ···················· |
| J | 7739 | hCoV-19/England/ALDP-95A6C0/2020\|EPI_ISL_553426\|20 | ···················· | ···················· | ····················· | ···················· | ···················· | ···················· |
| J | 7740 | hCoV-19/England/ALDP-954E46/2020\|EPI_ISL_553428\|20 | ···················· | ···················· | ····················· | ···················· | ···················· | ···················· |
| J | 7741 | hCoV-19/England/ALDP-95A37E/2020\|EPI_ISL_553429\|20 | ···················· | ···················· | ····················· | ···················· | ···················· | ···················· |
| J | 7742 | hCoV-19/England/ALDP-955513/2020\|EPI_ISL_553430\|20 | ···················· | ···················· | ····················· | ···················· | ···················· | ···················· |
| J | 7743 | hCoV-19/England/ALDP-954B8B/2020\|EPI_ISL_553431\|20 | ···················· | ···················· | ····················· | ···················· | ···················· | ···················· |
| J | 7744 | hCoV-19/England/ALDP-954BD6/2020\|EPI_ISL_553432\|20 | ···················· | ·A·················· | ····················· | ···················· | ···················· | ···················· |
| J | 7745 | hCoV-19/England/ALDP-95A675/2020\|EPI_ISL_553433\|20 | ···················· | ···················· | ····················· | ···················· | ···················· | ···················· |
| J | 7746 | hCoV-19/England/ALDP-95C88C/2020\|EPI_ISL_553435\|20 | ···················· | ···················· | ····················· | ···················· | ···················· | ···················· |
| J | 7747 | hCoV-19/England/ALDP-9554CB/2020\|EPI_ISL_553436\|20 | ···················· | ···················· | ····················· | ···················· | ···················· | ···················· |
| J | 7748 | hCoV-19/England/ALDP-95A8BB/2020\|EPI_ISL_553437\|20 | ···················· | ···················· | ····················· | ···················· | ···················· | ···················· |
| J | 7749 | hCoV-19/England/ALDP-95A648/2020\|EPI_ISL_553438\|20 | ···················· | ···················· | ····················· | ···················· | ···················· | ···················· |
| J | 7750 | hCoV-19/England/ALDP-95A8CA/2020\|EPI_ISL_553439\|20 | ···················· | ···················· | ····················· | ···················· | ···················· | ···················· |
| J | 7751 | hCoV-19/England/ALDP-95A02C/2020\|EPI_ISL_553440\|20 | ···················· | ···················· | ····················· | ···················· | ···················· | ···················· |
| J | 7752 | hCoV-19/England/ALDP-95A569/2020\|EPI_ISL_553441\|20 | ···················· | ···················· | ····················· | ···················· | ···················· | ···················· |
| J | 7753 | hCoV-19/England/ALDP-954F9E/2020\|EPI_ISL_553442\|20 | ···················· | ···················· | ····················· | ···················· | ···················· | ···················· |
| J | 7754 | hCoV-19/England/ALDP-95A3C9/2020\|EPI_ISL_553443\|20 | ···················· | ···················· | ····················· | ···················· | ···················· | ···················· |

| | | | | | | | | |
|---|---|---|---|---|---|---|---|---|
| J | 7755 | hCoV-19/England/ALDP-954C5B/2020\|EPI_ISL_553444\|20 | ···················· | ···················· | ····················· | ···················· | ···················· | ···················· |
| J | 7756 | hCoV-19/England/ALDP-95557D/2020\|EPI_ISL_553445\|20 | ···················· | ···················· | ····················· | ···················· | ···················· | ···················· |
| J | 7757 | hCoV-19/England/ALDP-9551D3/2020\|EPI_ISL_553446\|20 | ···················· | ···················· | ····················· | ···················· | ···················· | ···················· |
| J | 7758 | hCoV-19/England/ALDP-954A9D/2020\|EPI_ISL_553448\|20 | ···················· | ···················· | ····················· | ···················· | ···················· | ···················· |
| J | 7759 | hCoV-19/England/ALDP-954EBF/2020\|EPI_ISL_553449\|20 | ···················· | ···················· | ····················· | ···················· | ···················· | ···················· |
| J | 7760 | hCoV-19/England/ALDP-95A657/2020\|EPI_ISL_553450\|20 | ···················· | ···················· | ····················· | ···················· | ···················· | ···················· |
| J | 7761 | hCoV-19/England/ALDP-954FCB/2020\|EPI_ISL_553451\|20 | ···················· | ···················· | ····················· | ···················· | ···················· | ···················· |
| J | 7762 | hCoV-19/England/ALDP-954F8F/2020\|EPI_ISL_553452\|20 | ···················· | ···················· | ····················· | ···················· | ···················· | ···················· |
| J | 7763 | hCoV-19/England/ALDP-954B6D/2020\|EPI_ISL_553456\|20 | ···················· | ···················· | ····················· | ···················· | ···················· | ···················· |
| J | 7764 | hCoV-19/England/ALDP-95D1AE/2020\|EPI_ISL_553457\|20 | ···················· | ···················· | ····················· | ···················· | ···················· | ···················· |
| J | 7765 | hCoV-19/England/ALDP-954F61/2020\|EPI_ISL_553459\|20 | ···················· | ···················· | ····················· | ···················· | ···················· | ···················· |
| J | 7766 | hCoV-19/England/ALDP-95D232/2020\|EPI_ISL_553460\|20 | ···················· | ···················· | ····················· | ···················· | ···················· | ···················· |
| J | 7767 | hCoV-19/England/ALDP-955276/2020\|EPI_ISL_553461\|20 | ···················· | ···················· | ····················· | ···················· | ···················· | ···················· |
| J | 7768 | hCoV-19/England/ALDP-954B4F/2020\|EPI_ISL_553462\|20 | ···················· | ·A·················· | ····················· | ···················· | ···················· | ···················· |
| J | 7769 | hCoV-19/England/ALDP-94D4A8/2020\|EPI_ISL_553463\|20 | ···················· | ···················· | ····················· | ···················· | ···················· | ···················· |
| J | 7770 | hCoV-19/England/ALDP-9554F8/2020\|EPI_ISL_553464\|20 | ···················· | ···················· | ····················· | ···················· | ···················· | ···················· |
| J | 7771 | hCoV-19/England/ALDP-955522/2020\|EPI_ISL_553465\|20 | ···················· | ···················· | ····················· | ···················· | ···················· | ···················· |
| J | 7772 | hCoV-19/England/ALDP-95571D/2020\|EPI_ISL_553467\|20 | ···················· | ···················· | ····················· | ···················· | ···················· | ···················· |
| J | 7773 | hCoV-19/England/ALDP-954B7C/2020\|EPI_ISL_553469\|20 | ···················· | ···················· | ····················· | ···················· | ···················· | ···················· |
| J | 7774 | hCoV-19/England/ALDP-954DFE/2020\|EPI_ISL_553470\|20 | ···················· | ···················· | ····················· | ···················· | ···················· | ···················· |
| J | 7775 | hCoV-19/England/ALDP-955504/2020\|EPI_ISL_553472\|20 | ···················· | ···················· | ····················· | ···················· | ···················· | ···················· |
| J | 7776 | hCoV-19/England/ALDP-95CA3B/2020\|EPI_ISL_553474\|20 | ···················· | ···················· | ····················· | ···················· | ···················· | ···················· |
| J | 7777 | hCoV-19/England/ALDP-95A00E/2020\|EPI_ISL_553475\|20 | ···················· | ···················· | ····················· | ···················· | ···················· | ···················· |
| J | 7778 | hCoV-19/England/ALDP-95A3E7/2020\|EPI_ISL_553476\|20 | ···················· | ···················· | ····················· | ···················· | ···················· | ···················· |
| J | 7779 | hCoV-19/England/ALDP-954E91/2020\|EPI_ISL_553477\|20 | ···················· | ···················· | ····················· | ···················· | ···················· | ···················· |
| J | 7780 | hCoV-19/England/MILK-958413/2020\|EPI_ISL_554700\|20 | ···················· | ···················· | ····················· | ···················· | ···················· | ···················· |
| J | 7781 | hCoV-19/England/MILK-9582DD/2020\|EPI_ISL_554701\|20 | ···················· | ·A·················· | ····················· | ···················· | ···················· | ···················· |
| J | 7782 | hCoV-19/England/ALDP-944131/2020\|EPI_ISL_554703\|20 | ···················· | ···················· | ····················· | ···················· | ···················· | ···················· |
| J | 7783 | hCoV-19/England/MILK-958370/2020\|EPI_ISL_554705\|20 | ···················· | ···················· | ····················· | ···················· | ···················· | ···················· |
| J | 7784 | hCoV-19/England/MILK-95896F/2020\|EPI_ISL_554706\|20 | ···················· | ···················· | ····················· | ···················· | ···················· | ···················· |
| J | 7785 | hCoV-19/England/MILK-9588CC/2020\|EPI_ISL_554707\|20 | ···················· | ···················· | ····················· | ···················· | ···················· | ···················· |
| J | 7786 | hCoV-19/England/ALDP-94426B/2020\|EPI_ISL_554708\|20 | ···················· | ···················· | ····················· | ···················· | ···················· | ···················· |
| J | 7787 | hCoV-19/England/ALDP-944061/2020\|EPI_ISL_554710\|20 | ···················· | ···················· | ····················· | ···················· | ···················· | ···················· |
| J | 7788 | hCoV-19/England/MILK-9581D0/2020\|EPI_ISL_554712\|20 | ···················· | ···················· | ····················· | ···················· | ···················· | ···················· |
| J | 7789 | hCoV-19/England/MILK-957CA3/2020\|EPI_ISL_554713\|20 | ···················· | ···················· | ····················· | ···················· | ···················· | ···················· |
| J | 7790 | hCoV-19/England/MILK-9580B5/2020\|EPI_ISL_554714\|20 | ···················· | ···················· | ····················· | ···················· | ···················· | ···················· |
| J | 7791 | hCoV-19/England/MILK-95820A/2020\|EPI_ISL_554715\|20 | ···················· | ···················· | ····················· | ···················· | ···················· | ···················· |
| J | 7792 | hCoV-19/England/ALDP-944456/2020\|EPI_ISL_554716\|20 | ···················· | ···················· | ····················· | ···················· | ···················· | ···················· |
| J | 7793 | hCoV-19/England/MILK-9588F9/2020\|EPI_ISL_554717\|20 | ···················· | ···················· | ····················· | ···················· | ···················· | ···················· |
| J | 7794 | hCoV-19/England/MILK-958783/2020\|EPI_ISL_554718\|20 | ···················· | ···················· | ····················· | ···················· | ···················· | ···················· |
| J | 7795 | hCoV-19/England/MILK-958246/2020\|EPI_ISL_554719\|20 | ···················· | ···················· | ····················· | ···················· | ···················· | ···················· |
| J | 7796 | hCoV-19/England/MILK-958282/2020\|EPI_ISL_554720\|20 | ···················· | ···················· | ····················· | ···················· | ···················· | ···················· |
| J | 7797 | hCoV-19/England/ALDP-9442F2/2020\|EPI_ISL_554735\|20 | ···················· | ···················· | ····················· | ···················· | ···················· | ···················· |
| J | 7798 | hCoV-19/England/ALDP-944447/2020\|EPI_ISL_554743\|20 | ···················· | ···················· | ····················· | ···················· | ···················· | ···················· |
| J | 7799 | hCoV-19/England/MILK-9581A3/2020\|EPI_ISL_554744\|20 | ···················· | ·A·················· | ····················· | ···················· | ···················· | ···················· |
| J | 7800 | hCoV-19/England/MILK-9582BF/2020\|EPI_ISL_554745\|20 | ···················· | ···················· | ····················· | ···················· | ···················· | ···················· |
| J | 7801 | hCoV-19/England/MILK-95852F/2020\|EPI_ISL_554746\|20 | ···················· | ···················· | ····················· | ···················· | ···················· | ···················· |
| J | 7802 | hCoV-19/Wales/MILK-957E61/2020\|EPI_ISL_554747\|2020 | ···················· | ···················· | ····················· | ···················· | ···················· | ···················· |
| J | 7803 | hCoV-19/England/MILK-958404/2020\|EPI_ISL_554749\|20 | ···················· | ···················· | ····················· | ···················· | ···················· | ···················· |
| J | 7804 | hCoV-19/England/MILK-957FF5/2020\|EPI_ISL_554751\|20 | ···················· | ···················· | ····················· | ···················· | ···················· | ···················· |
| J | 7805 | hCoV-19/England/MILK-9583AD/2020\|EPI_ISL_554752\|20 | ···················· | ···················· | ····················· | ···················· | ···················· | ···················· |
| J | 7806 | hCoV-19/England/MILK-9585D4/2020\|EPI_ISL_554753\|20 | ···················· | ···················· | ····················· | ···················· | ···················· | ···················· |
| J | 7807 | hCoV-19/England/ALDP-944438/2020\|EPI_ISL_554754\|20 | ···················· | ···················· | ····················· | ···················· | ···················· | ···················· |
| J | 7808 | hCoV-19/England/ALDP-9443B3/2020\|EPI_ISL_554755\|20 | ···················· | ···················· | ····················· | ···················· | ···················· | ···················· |
| J | 7809 | hCoV-19/England/ALDP-944289/2020\|EPI_ISL_554756\|20 | ···················· | ···················· | ····················· | ···················· | ···················· | ···················· |
| J | 7810 | hCoV-19/England/ALDP-9442C5/2020\|EPI_ISL_554757\|20 | ···················· | ···················· | ····················· | ···················· | ···················· | ···················· |
| J | 7811 | hCoV-19/England/ALDP-9443A4/2020\|EPI_ISL_554759\|20 | ···················· | ···················· | ····················· | ···················· | ···················· | ···················· |
| J | 7812 | hCoV-19/England/MILK-95847D/2020\|EPI_ISL_554760\|20 | ···················· | ···················· | ····················· | ···················· | ···················· | ···················· |
| J | 7813 | hCoV-19/England/MILK-957E25/2020\|EPI_ISL_554762\|20 | ···················· | ···················· | ····················· | ···················· | ···················· | ···················· |
| J | 7814 | hCoV-19/England/MILK-9588DB/2020\|EPI_ISL_554765\|20 | ···················· | ···················· | ····················· | ···················· | ···················· | ···················· |
| J | 7815 | hCoV-19/England/MILK-958880/2020\|EPI_ISL_554766\|20 | ···················· | ···················· | ····················· | ···················· | ···················· | ···················· |
| J | 7816 | hCoV-19/England/ALDP-943F74/2020\|EPI_ISL_554767\|20 | ···················· | ···················· | ····················· | ···················· | ···················· | ···················· |
| J | 7817 | hCoV-19/England/MILK-9586A4/2020\|EPI_ISL_554768\|20 | ···················· | ···················· | ····················· | ···················· | ···················· | ···················· |
| J | 7818 | hCoV-19/England/MILK-95998C/2020\|EPI_ISL_554769\|20 | ···················· | ···················· | ····················· | ···················· | ···················· | ···················· |
| J | 7819 | hCoV-19/England/MILK-9581B2/2020\|EPI_ISL_554770\|20 | ···················· | ···················· | ····················· | ···················· | ···················· | ···················· |
| J | 7820 | hCoV-19/England/ALDP-94485A/2020\|EPI_ISL_554771\|20 | ···················· | ···················· | ····················· | ···················· | ···················· | ···················· |
| J | 7821 | hCoV-19/England/ALDP-944465/2020\|EPI_ISL_554772\|20 | ···················· | ···················· | ····················· | ···················· | ···················· | ···················· |
| J | 7822 | hCoV-19/England/MILK-958853/2020\|EPI_ISL_554773\|20 | ···················· | ···················· | ····················· | ···················· | ···················· | ···················· |
| J | 7823 | hCoV-19/Wales/MILK-9586C2/2020\|EPI_ISL_554774\|2020 | ···················· | ···················· | ····················· | ···················· | ···················· | ···················· |
| J | 7824 | hCoV-19/England/MILK-95839E/2020\|EPI_ISL_554775\|20 | ···················· | ···················· | ····················· | ···················· | ···················· | ···················· |
| J | 7825 | hCoV-19/England/ALDP-944605/2020\|EPI_ISL_554776\|20 | ···················· | ···················· | ····················· | ···················· | ···················· | ···················· |
| J | 7826 | hCoV-19/England/ALDP-944201/2020\|EPI_ISL_554777\|20 | ···················· | ···················· | ····················· | ···················· | ···················· | ···················· |
| J | 7827 | hCoV-19/England/ALDP-9441E6/2020\|EPI_ISL_554778\|20 | ···················· | ···················· | ····················· | ···················· | ···················· | ···················· |
| J | 7828 | hCoV-19/England/ALDP-9443C2/2020\|EPI_ISL_554780\|20 | ···················· | ···················· | ····················· | ···················· | ···················· | ···················· |
| J | 7829 | hCoV-19/England/ALDP-9445EA/2020\|EPI_ISL_554782\|20 | ···················· | ···················· | ····················· | ···················· | ···················· | ···················· |
| J | 7830 | hCoV-19/Wales/MILK-958729/2020\|EPI_ISL_554783\|2020 | ···················· | ···················· | ····················· | ···················· | ···················· | ···················· |
| J | 7831 | hCoV-19/England/MILK-9586E0/2020\|EPI_ISL_554784\|20 | ···················· | ···················· | ····················· | ···················· | ···················· | ···················· |
| J | 7832 | hCoV-19/England/MILK-9586D1/2020\|EPI_ISL_554785\|20 | ···················· | ···················· | ····················· | ···················· | ···················· | ···················· |
| J | 7833 | hCoV-19/England/MILK-958A99/2020\|EPI_ISL_554786\|20 | ···················· | ···················· | ····················· | ···················· | ···················· | ···················· |
| J | 7834 | hCoV-19/England/MILK-958361/2020\|EPI_ISL_554787\|20 | ···················· | ···················· | ····················· | ···················· | ···················· | ···················· |

| | | | | | | | | |
|---|---|---|---|---|---|---|---|---|
| J | 7835 | hCoV-19/England/MILK-957D73/2020\|EPI_ISL_554788\|20 | .................... | .................... | ..................... | .................... | .................... | .................... |
| J | 7836 | hCoV-19/England/MILK-9588EA/2020\|EPI_ISL_554789\|20 | .................... | .................... | ..................... | .................... | .................... | .................... |
| J | 7837 | hCoV-19/England/MILK-9586B3/2020\|EPI_ISL_554790\|20 | .................... | .................... | ..................... | .................... | .................... | .................... |
| J | 7838 | hCoV-19/England/MILK-957F04/2020\|EPI_ISL_554791\|20 | .................... | .................... | ..................... | .................... | .................... | .................... |
| J | 7839 | hCoV-19/England/ALDP-944492/2020\|EPI_ISL_554792\|20 | .................... | .................... | ..................... | .................... | .................... | .................... |
| J | 7840 | hCoV-19/England/MILK-957F7D/2020\|EPI_ISL_554793\|20 | .................... | .................... | ..................... | .................... | .................... | .................... |
| J | 7841 | hCoV-19/England/ALDP-944650/2020\|EPI_ISL_554794\|20 | .................... | .................... | ..................... | .................... | .................... | .................... |
| J | 7842 | hCoV-19/England/ALDP-943F38/2020\|EPI_ISL_554795\|20 | .................... | .................... | ..................... | .................... | .................... | .................... |
| J | 7843 | hCoV-19/England/ALDP-9445F9/2020\|EPI_ISL_554797\|20 | .................... | .................... | ..................... | .................... | .................... | .................... |
| J | 7844 | hCoV-19/England/ALDP-943F0B/2020\|EPI_ISL_554799\|20 | .................... | .................... | ..................... | .................... | .................... | .................... |
| J | 7845 | hCoV-19/England/ALDP-64D827/2020\|EPI_ISL_558100\|20 | .................... | .................... | ..................... | .................... | .................... | .................... |
| J | 7846 | hCoV-19/England/ALDP-651899/2020\|EPI_ISL_558101\|20 | .................... | .................... | ..................... | .................... | .................... | .................... |
| J | 7847 | hCoV-19/England/ALDP-64CEBD/2020\|EPI_ISL_558102\|20 | .................... | .................... | ..................... | .................... | .................... | .................... |
| J | 7848 | hCoV-19/England/ALDP-66ABC1/2020\|EPI_ISL_558105\|20 | .................... | .................... | ..................... | .................... | .................... | .................... |
| J | 7849 | hCoV-19/England/ALDP-66DD6E/2020\|EPI_ISL_558108\|20 | .................... | .................... | ..................... | .................... | .................... | .................... |
| J | 7850 | hCoV-19/England/MILK-576ECA/2020\|EPI_ISL_558251\|20 | .................... | .................... | ..................... | .................... | .................... | .................... |
| J | 7851 | hCoV-19/England/MILK-563A62/2020\|EPI_ISL_558252\|20 | .................... | .................... | ..................... | .................... | .................... | .................... |
| J | 7852 | hCoV-19/England/MILK-559EFA/2020\|EPI_ISL_558253\|20 | .................... | .................... | ..................... | .................... | .................... | .................... |
| J | 7853 | hCoV-19/England/MILK-58FF34/2020\|EPI_ISL_558254\|20 | .................... | .................... | ..................... | .................... | .................... | .................... |
| J | 7854 | hCoV-19/England/MILK-59460A/2020\|EPI_ISL_558255\|20 | .................... | .................... | ..................... | .................... | .................... | .................... |
| J | 7855 | hCoV-19/England/ALDP-59AD0F/2020\|EPI_ISL_558256\|20 | .................... | .A.................. | ..................... | .................... | .................... | .................... |
| J | 7856 | hCoV-19/England/MILK-55A3DC/2020\|EPI_ISL_558257\|20 | .................... | .................... | ..................... | .................... | .................... | .................... |
| J | 7857 | hCoV-19/England/MILK-5605FB/2020\|EPI_ISL_558258\|20 | .................... | .................... | ..................... | .................... | .................... | .................... |
| J | 7858 | hCoV-19/England/ALDP-59A358/2020\|EPI_ISL_558259\|20 | .................... | .................... | ..................... | .................... | .................... | .................... |
| J | 7859 | hCoV-19/England/ALDP-59B5BB/2020\|EPI_ISL_558260\|20 | .................... | .................... | ..................... | .................... | .................... | .................... |
| J | 7860 | hCoV-19/England/MILK-5660D6/2020\|EPI_ISL_558262\|20 | .................... | .................... | ..................... | .................... | .................... | .................... |
| J | 7861 | hCoV-19/England/ALDP-59A4FB/2020\|EPI_ISL_558263\|20 | .................... | .................... | ..................... | .................... | .................... | .................... |
| J | 7862 | hCoV-19/England/MILK-57B223/2020\|EPI_ISL_558264\|20 | .................... | .................... | ..................... | .................... | .................... | .................... |
| J | 7863 | hCoV-19/England/MILK-562605/2020\|EPI_ISL_558265\|20 | .................... | .................... | ...........T......... | .................... | .................... | .................... |
| J | 7864 | hCoV-19/England/MILK-576F4F/2020\|EPI_ISL_558266\|20 | .................... | .................... | ..................... | .................... | .................... | .................... |
| J | 7865 | hCoV-19/England/ALDP-599BCA/2020\|EPI_ISL_558268\|20 | .................... | .................... | ..................... | .................... | .................... | .................... |
| J | 7866 | hCoV-19/England/ALDP-59B7E2/2020\|EPI_ISL_558269\|20 | .................... | .................... | ..................... | .................... | .................... | .................... |
| J | 7867 | hCoV-19/England/ALDP-59BC2F/2020\|EPI_ISL_558270\|20 | .................... | .................... | ..................... | .................... | .................... | .................... |
| J | 7868 | hCoV-19/England/MILK-562D5B/2020\|EPI_ISL_558272\|20 | .................... | .................... | ..................... | .................... | .................... | .................... |
| J | 7869 | hCoV-19/England/MILK-56B217/2020\|EPI_ISL_558273\|20 | .................... | .................... | ..................... | .................... | .................... | .................... |
| J | 7870 | hCoV-19/England/ALDP-59A8FF/2020\|EPI_ISL_558276\|20 | .................... | .................... | ..................... | .................... | .................... | .................... |
| J | 7871 | hCoV-19/England/ALDP-59A279/2020\|EPI_ISL_558277\|20 | .................... | .................... | ..................... | .................... | .................... | .................... |
| J | 7872 | hCoV-19/England/MILK-597CAB/2020\|EPI_ISL_558278\|20 | .................... | .................... | ..................... | .................... | .................... | .................... |
| J | 7873 | hCoV-19/England/MILK-569677/2020\|EPI_ISL_558279\|20 | .................... | .................... | ..................... | .................... | .................... | .................... |
| J | 7874 | hCoV-19/England/MILK-57901C/2020\|EPI_ISL_558281\|20 | .................... | .................... | ..................... | .................... | .................... | .................... |
| J | 7875 | hCoV-19/England/MILK-569D72/2020\|EPI_ISL_558284\|20 | .................... | .................... | ..................... | .................... | .................... | .................... |
| J | 7876 | hCoV-19/England/MILK-55F10A/2020\|EPI_ISL_558285\|20 | .................... | .................... | ..................... | .................... | .................... | .................... |
| J | 7877 | hCoV-19/England/MILK-55C52F/2020\|EPI_ISL_558286\|20 | .................... | .................... | ..................... | .................... | .................... | .................... |
| J | 7878 | hCoV-19/England/MILK-57AA86/2020\|EPI_ISL_558287\|20 | .................... | .................... | ..................... | .................... | .................... | .................... |
| J | 7879 | hCoV-19/England/ALDP-59BC01/2020\|EPI_ISL_558288\|20 | .................... | .................... | ..................... | .................... | .................... | .................... |
| J | 7880 | hCoV-19/England/ALDP-59CA7F/2020\|EPI_ISL_558289\|20 | .................... | .................... | ..................... | .................... | .................... | .................... |
| J | 7881 | hCoV-19/England/MILK-5630D9/2020\|EPI_ISL_558290\|20 | .................... | .................... | ..................... | .................... | .................... | .................... |
| J | 7882 | hCoV-19/England/ALDP-599A72/2020\|EPI_ISL_558291\|20 | .................... | .A.................. | ..................... | .................... | .................... | .................... |
| J | 7883 | hCoV-19/England/MILK-562EC1/2020\|EPI_ISL_558294\|20 | .................... | .A.................. | ..................... | .................... | .................... | .................... |
| J | 7884 | hCoV-19/England/MILK-56514D/2020\|EPI_ISL_558295\|20 | .................... | .................... | ..................... | .................... | .................... | .................... |
| J | 7885 | hCoV-19/Wales/MILK-5932DA/2020\|EPI_ISL_558296\|2020 | .................... | .................... | ..................... | .................... | .................... | .................... |
| J | 7886 | hCoV-19/England/MILK-5757FD/2020\|EPI_ISL_558297\|20 | .................... | .................... | ..................... | .................... | .................... | .................... |
| J | 7887 | hCoV-19/England/MILK-559E27/2020\|EPI_ISL_558298\|20 | .................... | .................... | ..................... | .................... | .................... | .................... |
| J | 7888 | hCoV-19/England/MILK-59718E/2020\|EPI_ISL_558299\|20 | .................... | .................... | ..................... | .................... | .................... | .................... |
| J | 7889 | hCoV-19/England/ALDP-49A1B9/2020\|EPI_ISL_559400\|20 | .................... | .................... | ..................... | .................... | .................... | .................... |
| J | 7890 | hCoV-19/England/ALDP-4A2DC9/2020\|EPI_ISL_559401\|20 | .................... | .................... | ..................... | .................... | .................... | .................... |
| J | 7891 | hCoV-19/England/ALDP-499369/2020\|EPI_ISL_559402\|20 | .................... | .................... | ..................... | .................... | .................... | .................... |
| J | 7892 | hCoV-19/England/ALDP-4A22AC/2020\|EPI_ISL_559403\|20 | .................... | .................... | ..................... | .................... | .................... | .................... |
| J | 7893 | hCoV-19/England/ALDP-49843A/2020\|EPI_ISL_559404\|20 | .................... | .................... | ..................... | .................... | .................... | .................... |
| J | 7894 | hCoV-19/England/ALDP-49969D/2020\|EPI_ISL_559405\|20 | .................... | .................... | ..................... | .................... | .................... | .................... |
| J | 7895 | hCoV-19/England/ALDP-4986DA/2020\|EPI_ISL_559407\|20 | .................... | .................... | ..................... | .................... | .................... | .................... |
| J | 7896 | hCoV-19/England/ALDP-497C15/2020\|EPI_ISL_559408\|20 | .................... | .................... | ..................... | .................... | .................... | .................... |
| J | 7897 | hCoV-19/England/ALDP-498A47/2020\|EPI_ISL_559409\|20 | .................... | .................... | ..................... | .................... | .................... | .................... |
| J | 7898 | hCoV-19/England/ALDP-4A29A7/2020\|EPI_ISL_559410\|20 | .................... | .................... | ..................... | .................... | .................... | .................... |
| J | 7899 | hCoV-19/England/ALDP-4A1EB8/2020\|EPI_ISL_559411\|20 | .................... | .................... | ..................... | .................... | .................... | .................... |
| J | 7900 | hCoV-19/England/ALDP-4A2BFC/2020\|EPI_ISL_559412\|20 | .................... | .................... | ..................... | .................... | .................... | .................... |
| J | 7901 | hCoV-19/England/ALDP-499615/2020\|EPI_ISL_559413\|20 | .................... | .................... | ..................... | .................... | .................... | .................... |
| J | 7902 | hCoV-19/England/ALDP-499DC5/2020\|EPI_ISL_559414\|20 | .................... | .................... | ..................... | .................... | .................... | .................... |
| J | 7903 | hCoV-19/England/ALDP-49A3FF/2020\|EPI_ISL_559415\|20 | .................... | .................... | ..................... | .................... | .................... | .................... |
| J | 7904 | hCoV-19/England/ALDP-49A650/2020\|EPI_ISL_559416\|20 | .................... | .................... | ..................... | .................... | .................... | .................... |
| J | 7905 | hCoV-19/England/ALDP-4985FB/2020\|EPI_ISL_559417\|20 | .................... | .................... | ..................... | .................... | .................... | .................... |
| J | 7906 | hCoV-19/England/ALDP-49A6AB/2020\|EPI_ISL_559419\|20 | .................... | .................... | ..................... | .................... | .................... | .................... |
| J | 7907 | hCoV-19/England/ALDP-49A465/2020\|EPI_ISL_559420\|20 | .................... | .................... | ..................... | .................... | .................... | .................... |
| J | 7908 | hCoV-19/England/ALDP-4A25FE/2020\|EPI_ISL_559429\|20 | .................... | .................... | ..................... | .................... | .................... | .................... |
| J | 7909 | hCoV-19/England/ALDP-497899/2020\|EPI_ISL_559430\|20 | .................... | .................... | ..................... | .................... | .................... | .................... |
| J | 7910 | hCoV-19/England/ALDP-4984B2/2020\|EPI_ISL_559431\|20 | .................... | .................... | ..................... | .................... | .................... | .................... |
| J | 7911 | hCoV-19/England/ALDP-4996AC/2020\|EPI_ISL_559432\|20 | .................... | .................... | ..................... | .................... | .................... | .................... |
| J | 7912 | hCoV-19/England/ALDP-499B9E/2020\|EPI_ISL_559433\|20 | .................... | .................... | ..................... | .................... | .................... | .................... |
| J | 7913 | hCoV-19/England/ALDP-49878C/2020\|EPI_ISL_559434\|20 | .................... | .................... | ..................... | .................... | .................... | .................... |
| J | 7914 | hCoV-19/England/ALDP-4ACFE7/2020\|EPI_ISL_559436\|20 | .................... | .................... | ..................... | .................... | .................... | .................... |

| | | | | | | | |
|---|---|---|---|---|---|---|---|
| J | 7915 hCoV-19/England/ALDP-4A3539/2020\|EPI_ISL_559437\|20 | .................... | .................... | ..................... | .................... | .................... | .................... |
| J | 7916 hCoV-19/England/ALDP-4A875C/2020\|EPI_ISL_559438\|20 | .................... | .................... | ..................... | .................... | .................... | .................... |
| J | 7917 hCoV-19/England/ALDP-4AD9CA/2020\|EPI_ISL_559439\|20 | .................... | .................... | ..................... | .................... | .................... | .................... |
| J | 7918 hCoV-19/England/ALDP-4A7C21/2020\|EPI_ISL_559580\|20 | .................... | .................... | ..................... | .................... | .................... | .................... |
| J | 7919 hCoV-19/England/ALDP-4AA981/2020\|EPI_ISL_559581\|20 | .................... | .................... | ..................... | .................... | .................... | .................... |
| J | 7920 hCoV-19/England/ALDP-4A7C12/2020\|EPI_ISL_559582\|20 | .................... | .................... | ..................... | .................... | .................... | .................... |
| J | 7921 hCoV-19/England/ALDP-4A483F/2020\|EPI_ISL_559583\|20 | .................... | .................... | ..................... | .................... | .................... | .................... |
| J | 7922 hCoV-19/England/ALDP-4A6721/2020\|EPI_ISL_559584\|20 | .................... | .................... | ..................... | .................... | .................... | .................... |
| J | 7923 hCoV-19/England/ALDP-4A389A/2020\|EPI_ISL_559585\|20 | .................... | .................... | ..................... | .................... | .................... | .................... |
| J | 7924 hCoV-19/England/ALDP-49EC3C/2020\|EPI_ISL_559586\|20 | .................... | .................... | ..................... | .................... | .................... | .................... |
| J | 7925 hCoV-19/England/ALDP-4A776C/2020\|EPI_ISL_559587\|20 | .................... | .................... | ..................... | .................... | .................... | .................... |
| J | 7926 hCoV-19/England/ALDP-4A9551/2020\|EPI_ISL_559588\|20 | .................... | .................... | ..................... | .................... | .................... | .................... |
| J | 7927 hCoV-19/England/ALDP-4A924B/2020\|EPI_ISL_559589\|20 | .................... | .................... | ..................... | .................... | .................... | .................... |
| J | 7928 hCoV-19/England/ALDP-49E20D/2020\|EPI_ISL_559590\|20 | .................... | .................... | ..................... | .................... | .................... | .................... |
| J | 7929 hCoV-19/England/ALDP-4ADBB5/2020\|EPI_ISL_559592\|20 | .................... | .................... | ..................... | .................... | .................... | .................... |
| J | 7930 hCoV-19/England/ALDP-4AAC1F/2020\|EPI_ISL_559593\|20 | .................... | .................... | ..................... | .................... | ...T................ | .................... |
| J | 7931 hCoV-19/England/ALDP-4AE158/2020\|EPI_ISL_559594\|20 | .................... | .................... | ..................... | .................... | .................... | .................... |
| J | 7932 hCoV-19/England/ALDP-497DE5/2020\|EPI_ISL_559595\|20 | .................... | .................... | ..................... | .................... | .................... | .................... |
| J | 7933 hCoV-19/England/ALDP-4A7C03/2020\|EPI_ISL_559596\|20 | .................... | .................... | ..................... | .................... | .................... | .................... |
| J | 7934 hCoV-19/England/ALDP-4ADFAA/2020\|EPI_ISL_559597\|20 | .................... | .................... | ..................... | .................... | .................... | .................... |
| J | 7935 hCoV-19/England/ALDP-4A6BCB/2020\|EPI_ISL_559598\|20 | .................... | .................... | ..................... | .................... | .................... | .................... |
| J | 7936 hCoV-19/England/ALDP-4A82B5/2020\|EPI_ISL_559599\|20 | .................... | .................... | ..................... | .................... | .................... | .................... |
| J | 7937 hCoV-19/England/MILK-992F58/2020\|EPI_ISL_552000\|20 | .................... | .................... | ..................... | .................... | .................... | .................... |
| J | 7938 hCoV-19/England/CAMC-9915FD/2020\|EPI_ISL_552001\|20 | .................... | .................... | ..................... | .................... | .................... | .................... |
| J | 7939 hCoV-19/Wales/MILK-9928C6/2020\|EPI_ISL_552002\|2020 | .................... | .................... | ..................... | .................... | .................... | .................... |
| J | 7940 hCoV-19/Wales/MILK-992CCA/2020\|EPI_ISL_552003\|2020 | .................... | .................... | ..................... | .................... | .................... | .................... |
| J | 7941 hCoV-19/England/MILK-99331F/2020\|EPI_ISL_552011\|20 | .................... | .A.................. | ..................... | .................... | .................... | ..A................. |
| J | 7942 hCoV-19/England/MILK-993634/2020\|EPI_ISL_552012\|20 | .................... | .................... | ..................... | .................... | .................... | .................... |
| J | 7943 hCoV-19/England/MILK-970327/2020\|EPI_ISL_553236\|20 | .................... | .................... | ..................... | .................... | .................... | .................... |
| J | 7944 hCoV-19/England/MILK-9705B8/2020\|EPI_ISL_553237\|20 | .................... | .A.................. | ..................... | .................... | .................... | ..A................. |
| J | 7945 hCoV-19/England/MILK-970837/2020\|EPI_ISL_553238\|20 | .................... | .................... | ..................... | .................... | .................... | .................... |
| J | 7946 hCoV-19/England/MILK-970293/2020\|EPI_ISL_553243\|20 | .................... | .................... | ..................... | .................... | .................... | .................... |
| J | 7947 hCoV-19/England/MILK-97154E/2020\|EPI_ISL_553244\|20 | .................... | .................... | ..................... | .................... | .................... | .................... |
| J | 7948 hCoV-19/England/MILK-970DB0/2020\|EPI_ISL_553245\|20 | .................... | .................... | ..................... | .................... | .................... | .................... |
| J | 7949 hCoV-19/England/MILK-971669/2020\|EPI_ISL_553249\|20 | .................... | .................... | ..................... | .................... | .................... | .................... |
| J | 7950 hCoV-19/England/MILK-971362/2020\|EPI_ISL_553253\|20 | .................... | .................... | ..................... | .................... | .................... | .................... |
| J | 7951 hCoV-19/England/MILK-9709CB/2020\|EPI_ISL_553254\|20 | .................... | .................... | ..................... | .................... | .................... | ...............A.... |
| J | 7952 hCoV-19/England/MILK-970FC9/2020\|EPI_ISL_553255\|20 | .................... | .................... | ..................... | .................... | .................... | .................... |
| J | 7953 hCoV-19/England/MILK-970D74/2020\|EPI_ISL_553256\|20 | .................... | .................... | ..................... | .................... | .................... | .................... |
| J | 7954 hCoV-19/England/MILK-970AB9/2020\|EPI_ISL_553257\|20 | .................... | .................... | ..................... | .................... | .................... | .................... |
| J | 7955 hCoV-19/England/MILK-97061F/2020\|EPI_ISL_553258\|20 | .................... | .................... | ..................... | .................... | .................... | .................... |
| J | 7956 hCoV-19/England/MILK-970688/2020\|EPI_ISL_553259\|20 | .................... | .................... | ..................... | .................... | .................... | .................... |
| J | 7957 hCoV-19/Wales/MILK-9715C6/2020\|EPI_ISL_553260\|2020 | .................... | .................... | ..................... | .................... | .................... | .................... |
| J | 7958 hCoV-19/England/MILK-970E17/2020\|EPI_ISL_553261\|20 | .................... | .................... | ..................... | .................... | .................... | .................... |
| J | 7959 hCoV-19/England/MILK-970A40/2020\|EPI_ISL_553262\|20 | .................... | .................... | ..................... | .................... | .................... | .................... |
| J | 7960 hCoV-19/England/MILK-971335/2020\|EPI_ISL_553263\|20 | .................... | .................... | ..................... | .................... | .................... | .................... |
| J | 7961 hCoV-19/England/MILK-9708DD/2020\|EPI_ISL_553264\|20 | .................... | .................... | ..................... | .................... | .................... | .................... |
| J | 7962 hCoV-19/England/MILK-9701E1/2020\|EPI_ISL_553265\|20 | .................... | .................... | ..................... | .................... | .................... | .................... |
| J | 7963 hCoV-19/England/MILK-9707C1/2020\|EPI_ISL_553266\|20 | .................... | .................... | ..................... | .................... | .................... | .................... |
| J | 7964 hCoV-19/England/MILK-970ECC/2020\|EPI_ISL_553267\|20 | .................... | .................... | ..................... | .................... | .................... | .................... |
| J | 7965 hCoV-19/England/MILK-970521/2020\|EPI_ISL_553268\|20 | .................... | .................... | ..................... | .................... | .................... | .................... |
| J | 7966 hCoV-19/England/ALDP-91F919/2020\|EPI_ISL_553269\|20 | .................... | .................... | ..................... | .................... | .................... | .................... |
| J | 7967 hCoV-19/England/MILK-93CAC8/2020\|EPI_ISL_553270\|20 | .................... | .................... | ..................... | .................... | .................... | .................... |
| J | 7968 hCoV-19/England/ALDP-91F621/2020\|EPI_ISL_553271\|20 | .................... | .................... | ..................... | .................... | .................... | .................... |
| J | 7969 hCoV-19/England/ALDP-922193/2020\|EPI_ISL_553273\|20 | .................... | .................... | ..................... | .................... | .................... | .................... |
| J | 7970 hCoV-19/England/ALDP-91E65F/2020\|EPI_ISL_553274\|20 | .................... | .................... | ..................... | .................... | .................... | .................... |
| J | 7971 hCoV-19/England/MILK-93E413/2020\|EPI_ISL_553275\|20 | .................... | .................... | ..................... | .................... | .................... | .................... |
| J | 7972 hCoV-19/England/MILK-93C9F8/2020\|EPI_ISL_553276\|20 | .................... | .................... | ..................... | .................... | .................... | .................... |
| J | 7973 hCoV-19/England/ALDP-91F76A/2020\|EPI_ISL_553277\|20 | .................... | .................... | ..................... | .................... | .................... | .................... |
| J | 7974 hCoV-19/England/ALDP-92239D/2020\|EPI_ISL_553278\|20 | .................... | .................... | ..................... | .................... | .................... | .................... |
| J | 7975 hCoV-19/England/ALDP-91F7C4/2020\|EPI_ISL_553280\|20 | .................... | .................... | ..................... | .................... | .................... | .................... |
| J | 7976 hCoV-19/England/ALDP-9221DF/2020\|EPI_ISL_553281\|20 | .................... | .................... | ..................... | .................... | .................... | .................... |
| J | 7977 hCoV-19/England/ALDP-922FA8/2020\|EPI_ISL_553282\|20 | .................... | .................... | ..................... | .................... | .................... | .................... |
| J | 7978 hCoV-19/England/ALDP-921D72/2020\|EPI_ISL_553283\|20 | .................... | .................... | ..................... | .................... | .................... | .................... |
| J | 7979 hCoV-19/England/ALDP-91C6E8/2020\|EPI_ISL_553284\|20 | .................... | .................... | ..................... | .................... | .................... | .................... |
| J | 7980 hCoV-19/England/ALDP-91E90B/2020\|EPI_ISL_553285\|20 | .................... | .R.................. | ..................... | .................... | .................... | .................... |
| J | 7981 hCoV-19/England/ALDP-921D09/2020\|EPI_ISL_553286\|20 | .................... | .................... | ..................... | .................... | .................... | .................... |
| J | 7982 hCoV-19/England/ALDP-90BBFC/2020\|EPI_ISL_553287\|20 | .................... | .................... | ..................... | .................... | .................... | .................... |
| J | 7983 hCoV-19/England/ALDP-92248B/2020\|EPI_ISL_553288\|20 | .................... | .................... | ..................... | .................... | .................... | .................... |
| J | 7984 hCoV-19/England/ALDP-91E7C5/2020\|EPI_ISL_553289\|20 | .................... | .A.................. | ..................... | .................... | .................... | .................... |
| J | 7985 hCoV-19/England/ALDP-91F278/2020\|EPI_ISL_553290\|20 | .................... | .................... | ..................... | .................... | .................... | .................... |
| J | 7986 hCoV-19/England/ALDP-912F14/2020\|EPI_ISL_553291\|20 | .................... | .................... | ..................... | .................... | .................... | .................... |
| J | 7987 hCoV-19/England/ALDP-915139/2020\|EPI_ISL_553292\|20 | .................... | .................... | ..................... | .................... | .................... | .................... |
| J | 7988 hCoV-19/England/MILK-93C873/2020\|EPI_ISL_553295\|20 | .................... | .................... | ..................... | .................... | .................... | .................... |
| J | 7989 hCoV-19/England/ALDP-90CA0D/2020\|EPI_ISL_553297\|20 | .................... | .................... | ..................... | .................... | .................... | .................... |
| J | 7990 hCoV-19/England/ALDP-921E33/2020\|EPI_ISL_553298\|20 | .................... | .................... | ..................... | .................... | .................... | .................... |
| J | 7991 hCoV-19/England/ALDP-921FD6/2020\|EPI_ISL_553299\|20 | .................... | .................... | ..................... | .................... | .................... | .................... |
| J | 7992 hCoV-19/England/ALDP-91F64F/2020\|EPI_ISL_553300\|20 | .................... | .................... | ...........T......... | .................... | .................... | .................... |
| J | 7993 hCoV-19/England/ALDP-907264/2020\|EPI_ISL_553301\|20 | .................... | .................... | ..................... | .................... | .................... | .................... |
| J | 7994 hCoV-19/England/ALDP-90CAB2/2020\|EPI_ISL_553302\|20 | .................... | .................... | ..................... | .................... | .................... | .................... |

| | | | | | | | | |
|---|---|---|---|---|---|---|---|---|
| J | 7995 | hCoV-19/England/ALDP-9072FB/2020\|EPI_ISL_553303\|20 | ···················· | ···················· | ····················· | ···················· | ···················· | ···················· |
| J | 7996 | hCoV-19/England/ALDP-91E859/2020\|EPI_ISL_553305\|20 | ···················· | ·A·················· | ····················· | ···················· | ···················· | ···················· |
| J | 7997 | hCoV-19/England/ALDP-922CFC/2020\|EPI_ISL_553307\|20 | ···················· | ···················· | ····················· | ···················· | ···················· | ···················· |
| J | 7998 | hCoV-19/England/ALDP-9231B0/2020\|EPI_ISL_553308\|20 | ···················· | ···················· | ····················· | ···················· | ···················· | ···················· |
| J | 7999 | hCoV-19/England/ALDP-9230A4/2020\|EPI_ISL_553309\|20 | ···················· | ···················· | ····················· | ···················· | ···················· | ···················· |
| J | 8000 | hCoV-19/England/ALDP-91F8EF/2020\|EPI_ISL_553310\|20 | ···················· | ···················· | ····················· | ···················· | ···················· | ···················· |
| J | 8001 | hCoV-19/England/ALDP-91F542/2020\|EPI_ISL_553311\|20 | ···················· | ···················· | ····················· | ···················· | ···················· | ···················· |
| J | 8002 | hCoV-19/England/ALDP-921EF7/2020\|EPI_ISL_553312\|20 | ···················· | ···················· | ····················· | ···················· | ···················· | ···················· |
| J | 8003 | hCoV-19/England/ALDP-91E0E8/2020\|EPI_ISL_553313\|20 | ···················· | ···················· | ····················· | ···················· | ···················· | ···················· |
| J | 8004 | hCoV-19/England/ALDP-907343/2020\|EPI_ISL_553314\|20 | ···················· | ···················· | ····················· | ···················· | ···················· | ···················· |
| J | 8005 | hCoV-19/England/ALDP-922360/2020\|EPI_ISL_553315\|20 | ···················· | ···················· | ····················· | ···················· | ···················· | ···················· |
| J | 8006 | hCoV-19/England/ALDP-91F700/2020\|EPI_ISL_553316\|20 | ···················· | ···················· | ····················· | ···················· | ···················· | ···················· |
| J | 8007 | hCoV-19/England/ALDP-91F72E/2020\|EPI_ISL_553317\|20 | ···················· | ···················· | ····················· | ···················· | ···················· | ···················· |
| J | 8008 | hCoV-19/England/ALDP-91E4A0/2020\|EPI_ISL_553318\|20 | ···················· | ···················· | ····················· | ···················· | ···················· | ···················· |
| J | 8009 | hCoV-19/England/ALDP-921D45/2020\|EPI_ISL_553319\|20 | ···················· | ···················· | ····················· | ···················· | ···················· | ···················· |
| J | 8010 | hCoV-19/England/ALDP-91C90D/2020\|EPI_ISL_553321\|20 | ···················· | ·A·················· | ····················· | ···················· | ···················· | ···················· |
| J | 8011 | hCoV-19/England/ALDP-91AC51/2020\|EPI_ISL_553322\|20 | ···················· | ···················· | ····················· | ···················· | ···················· | ···················· |
| J | 8012 | hCoV-19/England/MILK-93E0A6/2020\|EPI_ISL_553323\|20 | ···················· | ···················· | ····················· | ···················· | ···················· | ···················· |
| J | 8013 | hCoV-19/England/ALDP-91E7D4/2020\|EPI_ISL_553325\|20 | ···················· | ···················· | ····················· | ···················· | ···················· | ···················· |
| J | 8014 | hCoV-19/England/ALDP-91A811/2020\|EPI_ISL_553326\|20 | ···················· | ···················· | ····················· | ···················· | ···················· | ···················· |
| J | 8015 | hCoV-19/England/ALDP-92460B/2020\|EPI_ISL_553328\|20 | ···················· | ···················· | ····················· | ···················· | ···················· | ···················· |
| J | 8016 | hCoV-19/England/ALDP-91F603/2020\|EPI_ISL_553329\|20 | ···················· | ···················· | ····················· | ···················· | ···················· | ···················· |
| J | 8017 | hCoV-19/Wales/ALDP-90C77F/2020\|EPI_ISL_553330\|2020 | ···················· | ···················· | ····················· | ···················· | ···················· | ···················· |
| J | 8018 | hCoV-19/England/ALDP-924692/2020\|EPI_ISL_553331\|20 | ···················· | ···················· | ····················· | ···················· | ···················· | ···················· |
| J | 8019 | hCoV-19/England/ALDP-9151DF/2020\|EPI_ISL_553333\|20 | ···················· | ···················· | ····················· | ···················· | ···················· | ···················· |
| J | 8020 | hCoV-19/England/ALDP-921C93/2020\|EPI_ISL_553334\|20 | ···················· | ·A·················· | ····················· | ···················· | ···················· | ···················· |
| J | 8021 | hCoV-19/England/ALDP-923095/2020\|EPI_ISL_553344\|20 | ···················· | ···················· | ····················· | ···················· | ···················· | ···················· |
| J | 8022 | hCoV-19/England/ALDP-91CF29/2020\|EPI_ISL_553345\|20 | ···················· | ···················· | ····················· | ···················· | ···················· | ···················· |
| J | 8023 | hCoV-19/England/ALDP-95573B/2020\|EPI_ISL_554567\|20 | ···················· | ···················· | ····················· | ···················· | ···················· | ···················· |
| J | 8024 | hCoV-19/England/ALDP-9509D0/2020\|EPI_ISL_554568\|20 | ···················· | ···················· | ····················· | ···················· | ···················· | ···················· |
| J | 8025 | hCoV-19/England/ALDP-950A0A/2020\|EPI_ISL_554569\|20 | ···················· | ···················· | ····················· | ···················· | ···················· | ···················· |
| J | 8026 | hCoV-19/England/ALDP-950D3E/2020\|EPI_ISL_554570\|20 | ···················· | ···················· | ····················· | ···················· | ···················· | ···················· |
| J | 8027 | hCoV-19/England/ALDP-9536A9/2020\|EPI_ISL_554571\|20 | ···················· | ···················· | ····················· | ···················· | ···················· | ···················· |
| J | 8028 | hCoV-19/England/ALDP-950CE6/2020\|EPI_ISL_554572\|20 | ···················· | ···················· | ····················· | ···················· | ···················· | ···················· |
| J | 8029 | hCoV-19/England/ALDP-951517/2020\|EPI_ISL_554573\|20 | ···················· | ···················· | ····················· | ···················· | ···················· | ···················· |
| J | 8030 | hCoV-19/England/ALDP-9508B5/2020\|EPI_ISL_554575\|20 | ···················· | ···················· | ····················· | ···················· | ···················· | ···················· |
| J | 8031 | hCoV-19/England/ALDP-9514CF/2020\|EPI_ISL_554576\|20 | ···················· | ···················· | ····················· | ···················· | ···················· | ···················· |
| J | 8032 | hCoV-19/England/ALDP-95125C/2020\|EPI_ISL_554577\|20 | ···················· | ·A·················· | ····················· | ···················· | ···················· | ···················· |
| J | 8033 | hCoV-19/England/ALDP-950B43/2020\|EPI_ISL_554578\|20 | ···················· | ···················· | ····················· | ···················· | ···················· | ···················· |
| J | 8034 | hCoV-19/England/ALDP-953E1A/2020\|EPI_ISL_554579\|20 | ···················· | ···················· | ····················· | ···················· | ···················· | ···················· |
| J | 8035 | hCoV-19/England/ALDP-951456/2020\|EPI_ISL_554580\|20 | ···················· | ···················· | ····················· | ···················· | ···················· | ···················· |
| J | 8036 | hCoV-19/England/ALDP-950BAD/2020\|EPI_ISL_554582\|20 | ···················· | ···················· | ····················· | ···················· | ···················· | ···················· |
| J | 8037 | hCoV-19/England/ALDP-95314E/2020\|EPI_ISL_554583\|20 | ···················· | ···················· | ····················· | ···················· | ···················· | ···················· |
| J | 8038 | hCoV-19/England/ALDP-955980/2020\|EPI_ISL_554585\|20 | ···················· | ···················· | ····················· | ···················· | ···················· | ···················· |
| J | 8039 | hCoV-19/England/ALDP-9513FF/2020\|EPI_ISL_554586\|20 | ···················· | ···················· | ····················· | ···················· | ···················· | ···················· |
| J | 8040 | hCoV-19/England/ALDP-950CC8/2020\|EPI_ISL_554587\|20 | ···················· | ···················· | ····················· | ···················· | ···················· | ···················· |
| J | 8041 | hCoV-19/England/ALDP-9508F1/2020\|EPI_ISL_554588\|20 | ···················· | ···················· | ····················· | ···················· | ···················· | ···················· |
| J | 8042 | hCoV-19/England/ALDP-954286/2020\|EPI_ISL_554589\|20 | ···················· | ···················· | ····················· | ···················· | ···················· | ···················· |
| J | 8043 | hCoV-19/England/ALDP-950D89/2020\|EPI_ISL_554590\|20 | ···················· | ···················· | ····················· | ···················· | ···················· | ···················· |
| J | 8044 | hCoV-19/England/ALDP-950879/2020\|EPI_ISL_554592\|20 | ···················· | ···················· | ····················· | ···················· | ···················· | ···················· |
| J | 8045 | hCoV-19/England/ALDP-950CB9/2020\|EPI_ISL_554594\|20 | ···················· | ···················· | ····················· | ···················· | ···················· | ···················· |
| J | 8046 | hCoV-19/England/ALDP-9554AD/2020\|EPI_ISL_554595\|20 | ···················· | ···················· | ····················· | ···················· | ···················· | ···················· |
| J | 8047 | hCoV-19/England/ALDP-950E2C/2020\|EPI_ISL_554596\|20 | ···················· | ···················· | ····················· | ···················· | ···················· | ···················· |
| J | 8048 | hCoV-19/England/ALDP-951562/2020\|EPI_ISL_554597\|20 | ···················· | ···················· | ····················· | ···················· | ···················· | ···················· |
| J | 8049 | hCoV-19/England/ALDP-9533DF/2020\|EPI_ISL_554598\|20 | ···················· | ···················· | ····················· | ···················· | ···················· | ···················· |
| J | 8050 | hCoV-19/England/ALDP-950B52/2020\|EPI_ISL_554599\|20 | ···················· | ·A·················· | ····················· | ···················· | ···················· | ···················· |
| J | 8051 | hCoV-19/England/ALDP-95364F/2020\|EPI_ISL_554600\|20 | ···················· | ···················· | ····················· | ···················· | ···················· | ···················· |
| J | 8052 | hCoV-19/England/ALDP-95079A/2020\|EPI_ISL_554602\|20 | ···················· | ···················· | ····················· | ···················· | ···················· | ···················· |
| J | 8053 | hCoV-19/England/ALDP-95078B/2020\|EPI_ISL_554603\|20 | ···················· | ···················· | ····················· | ···················· | ···················· | ···················· |
| J | 8054 | hCoV-19/England/ALDP-955944/2020\|EPI_ISL_554604\|20 | ···················· | ···················· | ····················· | ···················· | ···················· | ···················· |
| J | 8055 | hCoV-19/England/ALDP-9540B9/2020\|EPI_ISL_554605\|20 | ···················· | ···················· | ····················· | ···················· | ···················· | ···················· |
| J | 8056 | hCoV-19/England/ALDP-953FBD/2020\|EPI_ISL_554606\|20 | ···················· | ···················· | ····················· | ···················· | ···················· | ···················· |
| J | 8057 | hCoV-19/England/ALDP-953005/2020\|EPI_ISL_554607\|20 | ···················· | ···················· | ····················· | ···················· | ···················· | ···················· |
| J | 8058 | hCoV-19/England/ALDP-955407/2020\|EPI_ISL_554608\|20 | ···················· | ···················· | ····················· | ···················· | ···················· | ···················· |
| J | 8059 | hCoV-19/England/ALDP-950BE9/2020\|EPI_ISL_554609\|20 | ···················· | ···················· | ····················· | ···················· | ···················· | ···················· |
| J | 8060 | hCoV-19/England/ALDP-9540AA/2020\|EPI_ISL_554610\|20 | ···················· | ···················· | ····················· | ···················· | ···················· | ···················· |
| J | 8061 | hCoV-19/England/ALDP-9556C5/2020\|EPI_ISL_554612\|20 | ···················· | ···················· | ····················· | ···················· | ···················· | ···················· |
| J | 8062 | hCoV-19/England/ALDP-9532D2/2020\|EPI_ISL_554614\|20 | ···················· | ···················· | ····················· | ···················· | ···················· | ···················· |
| J | 8063 | hCoV-19/England/ALDP-943FCF/2020\|EPI_ISL_554615\|20 | ···················· | ···················· | ····················· | ···················· | ···················· | ···················· |
| J | 8064 | hCoV-19/England/ALDP-944395/2020\|EPI_ISL_554616\|20 | ···················· | ···················· | ····················· | ···················· | ···················· | ···················· |
| J | 8065 | hCoV-19/England/ALDP-94467E/2020\|EPI_ISL_554617\|20 | ···················· | ···················· | ····················· | ···················· | ···················· | ···················· |
| J | 8066 | hCoV-19/England/ALDP-944508/2020\|EPI_ISL_554618\|20 | ···················· | ···················· | ····················· | ···················· | ···················· | ···················· |
| J | 8067 | hCoV-19/England/MILK-957DEC/2020\|EPI_ISL_554619\|20 | ···················· | ···················· | ····················· | ···················· | ···················· | ···················· |
| J | 8068 | hCoV-19/England/MILK-957CFE/2020\|EPI_ISL_554622\|20 | ···················· | ···················· | ····················· | ···················· | ···················· | ···················· |
| J | 8069 | hCoV-19/England/ALDP-944553/2020\|EPI_ISL_554623\|20 | ···················· | ···················· | ··T·················· | ···················· | ···················· | ···················· |
| J | 8070 | hCoV-19/England/ALDP-9441B9/2020\|EPI_ISL_554624\|20 | ···················· | ···················· | ····················· | ···················· | ···················· | ···················· |
| J | 8071 | hCoV-19/England/ALDP-943FED/2020\|EPI_ISL_554625\|20 | ···················· | ···················· | ····················· | ···················· | ···················· | ···················· |
| J | 8072 | hCoV-19/England/ALDP-943FA1/2020\|EPI_ISL_554626\|20 | ···················· | ···················· | ····················· | ···················· | ···················· | ···················· |
| J | 8073 | hCoV-19/England/MILK-9585E3/2020\|EPI_ISL_554628\|20 | ···················· | ···················· | ····················· | ···················· | ···················· | ···················· |
| J | 8074 | hCoV-19/England/MILK-958194/2020\|EPI_ISL_554629\|20 | ···················· | ···················· | ····················· | ···················· | ···················· | ···················· |

| | | | | | | | | |
|---|---|---|---|---|---|---|---|---|
| J | 8075 | hCoV-19/England/ALDP-9444ED/2020\|EPI_ISL_554630\|20 | ···················· | ···················· | ····················· | ···················· | ···················· | ···················· |
| J | 8076 | hCoV-19/Wales/MILK-958862/2020\|EPI_ISL_554633\|2020 | ···················· | ···················· | ····················· | ···················· | ···················· | ···················· |
| J | 8077 | hCoV-19/England/MILK-957EAD/2020\|EPI_ISL_554635\|20 | ···················· | ···················· | ····················· | ···················· | ···················· | ···················· |
| J | 8078 | hCoV-19/England/MILK-957DCE/2020\|EPI_ISL_554636\|20 | ···················· | ···················· | ····················· | ···················· | ···················· | ···················· |
| J | 8079 | hCoV-19/England/MILK-95812B/2020\|EPI_ISL_554637\|20 | ···················· | ···················· | ····················· | ···················· | ···················· | ···················· |
| J | 8080 | hCoV-19/England/MILK-9588AE/2020\|EPI_ISL_554639\|20 | ···················· | ···················· | ····················· | ···················· | ···················· | ···················· |
| J | 8081 | hCoV-19/England/ALDP-9446BA/2020\|EPI_ISL_554640\|20 | ···················· | ···················· | ····················· | ···················· | ···················· | ···················· |
| J | 8082 | hCoV-19/England/MILK-95853E/2020\|EPI_ISL_554641\|20 | ···················· | ···················· | ····················· | ···················· | ···················· | ···················· |
| J | 8083 | hCoV-19/England/MILK-958167/2020\|EPI_ISL_554643\|20 | ···················· | ···················· | ····················· | ···················· | ···················· | ···················· |
| J | 8084 | hCoV-19/England/MILK-95805B/2020\|EPI_ISL_554644\|20 | ···················· | ···················· | ····················· | ···················· | ···················· | ···················· |
| J | 8085 | hCoV-19/England/MILK-957F9B/2020\|EPI_ISL_554647\|20 | ···················· | ···················· | ····················· | ···················· | ···················· | ···················· |
| J | 8086 | hCoV-19/England/MILK-9583DA/2020\|EPI_ISL_554648\|20 | ···················· | ···················· | ····················· | ···················· | ···················· | ···················· |
| J | 8087 | hCoV-19/England/ALDP-9443D1/2020\|EPI_ISL_554651\|20 | ···················· | ···················· | ····················· | ···················· | ···················· | ···················· |
| J | 8088 | hCoV-19/England/ALDP-944104/2020\|EPI_ISL_554656\|20 | ···················· | ···················· | ····················· | ···················· | ···················· | ···················· |
| J | 8089 | hCoV-19/England/ALDP-944614/2020\|EPI_ISL_554657\|20 | ···················· | ···················· | ····················· | ···················· | ···················· | ···················· |
| J | 8090 | hCoV-19/England/MILK-958219/2020\|EPI_ISL_554658\|20 | ···················· | ···················· | ····················· | ···················· | ···················· | ···················· |
| J | 8091 | hCoV-19/England/MILK-958774/2020\|EPI_ISL_554659\|20 | ···················· | ···················· | ····················· | ···················· | ···················· | ···················· |
| J | 8092 | hCoV-19/England/MILK-95897E/2020\|EPI_ISL_554660\|20 | ···················· | ···················· | ····················· | ···················· | ···················· | ···················· |
| J | 8093 | hCoV-19/England/ALDP-944544/2020\|EPI_ISL_554664\|20 | ···················· | ···················· | ····················· | ···················· | ···················· | ···················· |
| J | 8094 | hCoV-19/England/MILK-957C85/2020\|EPI_ISL_554665\|20 | ···················· | ···················· | ····················· | ···················· | ···················· | ···················· |
| J | 8095 | hCoV-19/England/MILK-957CB2/2020\|EPI_ISL_554675\|20 | ···················· | ···················· | ····················· | ···················· | ···················· | ···················· |
| J | 8096 | hCoV-19/England/ALDP-94CDCC/2020\|EPI_ISL_555898\|20 | ···················· | ···················· | ····················· | ···················· | ···················· | ···················· |
| J | 8097 | hCoV-19/England/ALDP-94CF11/2020\|EPI_ISL_555899\|20 | ···················· | ···················· | ····················· | ···················· | ···················· | ···················· |
| J | 8098 | hCoV-19/England/ALDP-94C7CE/2020\|EPI_ISL_555932\|20 | ···················· | ···················· | ····················· | ···················· | ···················· | ···················· |
| J | 8099 | hCoV-19/England/ALDP-94D46C/2020\|EPI_ISL_555933\|20 | ···················· | ···················· | ····················· | ···················· | ···················· | ···················· |
| J | 8100 | hCoV-19/England/ALDP-94F497/2020\|EPI_ISL_555934\|20 | ···················· | ···················· | ····················· | ···················· | ···················· | ···················· |
| J | 8101 | hCoV-19/England/ALDP-94C52E/2020\|EPI_ISL_555935\|20 | ···················· | ···················· | ····················· | ···················· | ···················· | ···················· |
| J | 8102 | hCoV-19/England/ALDP-94CD71/2020\|EPI_ISL_555936\|20 | ···················· | ···················· | ····················· | ···················· | ···················· | ···················· |
| J | 8103 | hCoV-19/England/ALDP-94CB2C/2020\|EPI_ISL_555937\|20 | ···················· | ···················· | ····················· | ···················· | ···················· | ···················· |
| J | 8104 | hCoV-19/England/ALDP-94CB59/2020\|EPI_ISL_555938\|20 | ···················· | ···················· | ····················· | ···················· | ···················· | ···················· |
| J | 8105 | hCoV-19/England/CAMC-949819/2020\|EPI_ISL_555939\|20 | ···················· | ·A·················· | ····················· | ···················· | ···················· | ···················· |
| J | 8106 | hCoV-19/England/ALDP-94CF8A/2020\|EPI_ISL_555941\|20 | ···················· | ···················· | ····················· | ···················· | ···················· | ···················· |
| J | 8107 | hCoV-19/England/ALDP-94C9E6/2020\|EPI_ISL_555942\|20 | ···················· | ···················· | ····················· | ···················· | ···················· | ···················· |
| J | 8108 | hCoV-19/England/ALDP-94C6A3/2020\|EPI_ISL_555943\|20 | ···················· | ···················· | ····················· | ···················· | ···················· | ···················· |
| J | 8109 | hCoV-19/England/ALDP-94C940/2020\|EPI_ISL_555944\|20 | ···················· | ···················· | ····················· | ···················· | ···················· | ···················· |
| J | 8110 | hCoV-19/England/CAMC-949239/2020\|EPI_ISL_555945\|20 | ···················· | ···················· | ····················· | ···················· | ···················· | ···················· |
| J | 8111 | hCoV-19/England/ALDP-94C719/2020\|EPI_ISL_555946\|20 | ···················· | ·A·················· | ····················· | ···················· | ···················· | ···················· |
| J | 8112 | hCoV-19/England/CAMC-94A308/2020\|EPI_ISL_555948\|20 | ···················· | ···················· | ····················· | ···················· | ···················· | ···················· |
| J | 8113 | hCoV-19/England/ALDP-94C430/2020\|EPI_ISL_555949\|20 | ···················· | ···················· | ····················· | ···················· | ···················· | ···················· |
| J | 8114 | hCoV-19/England/ALDP-94CDBD/2020\|EPI_ISL_555951\|20 | ···················· | ···················· | ····················· | ···················· | ···················· | ···················· |
| J | 8115 | hCoV-19/England/ALDP-94C96E/2020\|EPI_ISL_555952\|20 | ···················· | ···················· | ····················· | ···················· | ···················· | ···················· |
| J | 8116 | hCoV-19/England/ALDP-94CDF9/2020\|EPI_ISL_555954\|20 | ···················· | ···················· | ····················· | ···················· | ···················· | ···················· |
| J | 8117 | hCoV-19/England/CAMC-94A1C2/2020\|EPI_ISL_555955\|20 | ···················· | ·········A·········· | ····················· | ···················· | ···················· | ···················· |
| J | 8118 | hCoV-19/England/ALDP-94CDEA/2020\|EPI_ISL_555956\|20 | ···················· | ···················· | ····················· | ···················· | ···················· | ···················· |
| J | 8119 | hCoV-19/England/ALDP-94D0FF/2020\|EPI_ISL_555957\|20 | ···················· | ···················· | ····················· | ···················· | ···················· | ···················· |
| J | 8120 | hCoV-19/England/ALDP-94CD53/2020\|EPI_ISL_555958\|20 | ···················· | ···················· | ····················· | ···················· | ···················· | ···················· |
| J | 8121 | hCoV-19/England/ALDP-94D01D/2020\|EPI_ISL_555959\|20 | ···················· | ···················· | ····················· | ···················· | ···················· | ···················· |
| J | 8122 | hCoV-19/England/ALDP-94F691/2020\|EPI_ISL_555960\|20 | ···················· | ···················· | ····················· | ···················· | ···················· | ···················· |
| J | 8123 | hCoV-19/England/ALDP-94F136/2020\|EPI_ISL_555961\|20 | ···················· | ···················· | ····················· | ···················· | ···················· | ···················· |
| J | 8124 | hCoV-19/England/ALDP-94CCDE/2020\|EPI_ISL_555962\|20 | ···················· | ···················· | ····················· | ···················· | ···················· | ···················· |
| J | 8125 | hCoV-19/England/ALDP-94CC83/2020\|EPI_ISL_555963\|20 | ···················· | ···················· | ····················· | ···················· | ···················· | ···················· |
| J | 8126 | hCoV-19/England/ALDP-94CF20/2020\|EPI_ISL_555964\|20 | ···················· | ···················· | ····················· | ···················· | ···················· | ···················· |
| J | 8127 | hCoV-19/England/ALDP-94CBA4/2020\|EPI_ISL_555965\|20 | ···················· | ···················· | ····················· | ···················· | ···················· | ···················· |
| J | 8128 | hCoV-19/England/ALDP-94D11A/2020\|EPI_ISL_555966\|20 | ···················· | ···················· | ····················· | ···················· | ···················· | ···················· |
| J | 8129 | hCoV-19/England/ALDP-94C3F7/2020\|EPI_ISL_555968\|20 | ···················· | ···················· | ····················· | ···················· | ···················· | ···················· |
| J | 8130 | hCoV-19/England/ALDP-94F400/2020\|EPI_ISL_555969\|20 | ···················· | ···················· | ····················· | ···················· | ···················· | ···················· |
| J | 8131 | hCoV-19/England/ALDP-94C694/2020\|EPI_ISL_555970\|20 | ···················· | ···················· | ····················· | ···················· | ···················· | ···················· |
| J | 8132 | hCoV-19/England/ALDP-94F655/2020\|EPI_ISL_555971\|20 | ···················· | ···················· | ····················· | ···················· | ···················· | ···················· |
| J | 8133 | hCoV-19/England/ALDP-94F27F/2020\|EPI_ISL_555972\|20 | ···················· | ···················· | ····················· | ···················· | ···················· | ···················· |
| J | 8134 | hCoV-19/England/CAMC-94A2ED/2020\|EPI_ISL_555973\|20 | ···················· | ···················· | ····················· | ···················· | ···················· | ···················· |
| J | 8135 | hCoV-19/England/ALDP-94CFC6/2020\|EPI_ISL_555974\|20 | ···················· | ···················· | ····················· | ···················· | ···················· | ···················· |
| J | 8136 | hCoV-19/England/ALDP-94CEF6/2020\|EPI_ISL_555975\|20 | ···················· | ···················· | ····················· | ···················· | ···················· | ···················· |
| J | 8137 | hCoV-19/England/CAMC-94A1FF/2020\|EPI_ISL_555976\|20 | ···················· | ···················· | ····················· | ···················· | ···················· | ···················· |
| J | 8138 | hCoV-19/England/ALDP-94F38B/2020\|EPI_ISL_555977\|20 | ···················· | ···················· | ····················· | ···················· | ···················· | ···················· |
| J | 8139 | hCoV-19/Wales/ALDP-94C764/2020\|EPI_ISL_555978\|2020 | ···················· | ···················· | ····················· | ···················· | ···················· | ···················· |
| J | 8140 | hCoV-19/England/ALDP-94F2CA/2020\|EPI_ISL_555979\|20 | ···················· | ···················· | ····················· | ···················· | ···················· | ···················· |
| J | 8141 | hCoV-19/England/ALDP-94CA7A/2020\|EPI_ISL_555980\|20 | ···················· | ···················· | ····················· | ···················· | ···················· | ···················· |
| J | 8142 | hCoV-19/England/ALDP-94CCB0/2020\|EPI_ISL_555981\|20 | ···················· | ·A·················· | ····················· | ···················· | ···················· | ···················· |
| J | 8143 | hCoV-19/England/ALDP-94CEE7/2020\|EPI_ISL_555983\|20 | ···················· | ···················· | ····················· | ···················· | ···················· | ···················· |
| J | 8144 | hCoV-19/England/ALDP-94F2AC/2020\|EPI_ISL_555984\|20 | ···················· | ···················· | ····················· | ···················· | ···················· | ···················· |
| J | 8145 | hCoV-19/England/ALDP-94CD26/2020\|EPI_ISL_555985\|20 | ···················· | ···················· | ····················· | ···················· | ···················· | ···················· |
| J | 8146 | hCoV-19/England/ALDP-94C47C/2020\|EPI_ISL_555987\|20 | ···················· | ···················· | ····················· | ···················· | ···················· | ···················· |
| J | 8147 | hCoV-19/England/ALDP-94C99B/2020\|EPI_ISL_555988\|20 | ···················· | ···················· | ····················· | ···················· | ···················· | ···················· |
| J | 8148 | hCoV-19/England/ALDP-94F215/2020\|EPI_ISL_555989\|20 | ···················· | ···················· | ····················· | ···················· | ···················· | ···················· |
| J | 8149 | hCoV-19/England/ALDP-94C53D/2020\|EPI_ISL_555991\|20 | ···················· | ···················· | ····················· | ···················· | ···················· | ···················· |
| J | 8150 | hCoV-19/England/ALDP-94F664/2020\|EPI_ISL_555994\|20 | ···················· | ···················· | ····················· | ···················· | ···················· | ···················· |
| J | 8151 | hCoV-19/Wales/ALDP-94F594/2020\|EPI_ISL_555995\|2020 | ···················· | ···················· | ····················· | ···················· | ···················· | ···················· |
| J | 8152 | hCoV-19/England/ALDP-94C825/2020\|EPI_ISL_555996\|20 | ···················· | ···················· | ····················· | ···················· | ···················· | ···················· |
| J | 8153 | hCoV-19/England/ALDP-66ACDD/2020\|EPI_ISL_558110\|20 | ···················· | ···················· | ····················· | ···················· | ···················· | ···················· |
| J | 8154 | hCoV-19/England/ALDP-64A3A1/2020\|EPI_ISL_558112\|20 | ···················· | ···················· | ····················· | ···················· | ···················· | ···················· |

| | | | | | | | |
|---|---|---|---|---|---|---|---|
| J | 8155 hCoV-19/England/ALDP-647EFE/2020\|EPI_ISL_558114\|20 | ···················· | ···················· | ····················· | ···················· | ···················· | ···················· |
| J | 8156 hCoV-19/England/ALDP-647D88/2020\|EPI_ISL_558115\|20 | ···················· | ···················· | ····················· | ···················· | ···················· | ···················· |
| J | 8157 hCoV-19/England/ALDP-64B3EC/2020\|EPI_ISL_558116\|20 | ········Y··········· | ···················· | ····················· | ···················· | ···················· | ···················· |
| J | 8158 hCoV-19/England/ALDP-66EDE5/2020\|EPI_ISL_558118\|20 | ···················· | ···················· | ····················· | ···················· | ···················· | ···················· |
| J | 8159 hCoV-19/England/ALDP-66B331/2020\|EPI_ISL_558119\|20 | ···················· | ···················· | ····················· | ···················· | ···················· | ···················· |
| J | 8160 hCoV-19/England/ALDP-68C16D/2020\|EPI_ISL_558120\|20 | ···················· | ···················· | ····················· | ···················· | ···················· | ···················· |
| J | 8161 hCoV-19/England/ALDP-66D232/2020\|EPI_ISL_558121\|20 | ···················· | ···················· | ····················· | ···················· | ···················· | ···················· |
| J | 8162 hCoV-19/England/ALDP-65920C/2020\|EPI_ISL_558122\|20 | ···················· | ···················· | ····················· | ···················· | ···················· | ···················· |
| J | 8163 hCoV-19/England/ALDP-68E013/2020\|EPI_ISL_558123\|20 | ···················· | ···················· | ····················· | ···················· | ···················· | ···················· |
| J | 8164 hCoV-19/England/ALDP-64A30B/2020\|EPI_ISL_558124\|20 | ···················· | ···················· | ····················· | ···················· | ···················· | ···················· |
| J | 8165 hCoV-19/England/ALDP-658AF6/2020\|EPI_ISL_558125\|20 | ···················· | ···················· | ····················· | ···················· | ···················· | ···················· |
| J | 8166 hCoV-19/England/ALDP-68E602/2020\|EPI_ISL_558128\|20 | ···················· | ···················· | ····················· | ···················· | ···················· | ···················· |
| J | 8167 hCoV-19/England/ALDP-691235/2020\|EPI_ISL_558129\|20 | ···················· | ···················· | ····················· | ···················· | ···················· | ···················· |
| J | 8168 hCoV-19/England/ALDP-4994A2/2020\|EPI_ISL_559421\|20 | ···················· | ···················· | ····················· | ···················· | ···················· | ···················· |
| J | 8169 hCoV-19/England/ALDP-4A1D06/2020\|EPI_ISL_559422\|20 | ···················· | ···················· | ····················· | ···················· | ···················· | ···················· |
| J | 8170 hCoV-19/England/ALDP-497CAC/2020\|EPI_ISL_559423\|20 | ···················· | ···················· | ····················· | ···················· | ···················· | ···················· |
| J | 8171 hCoV-19/England/ALDP-4A30B0/2020\|EPI_ISL_559424\|20 | ···················· | ···················· | ····················· | ···················· | ···················· | ···················· |
| J | 8172 hCoV-19/England/ALDP-49A5AE/2020\|EPI_ISL_559425\|20 | ···················· | ···················· | ····················· | ···················· | ···················· | ···················· |
| J | 8173 hCoV-19/England/ALDP-4A20A2/2020\|EPI_ISL_559426\|20 | ···················· | ···················· | ····················· | ···················· | ···················· | ···················· |
| J | 8174 hCoV-19/England/ALDP-49A447/2020\|EPI_ISL_559428\|20 | ···················· | ···················· | ····················· | ···················· | ···················· | ···················· |
| J | 8175 hCoV-19/England/ALDP-4A5661/2020\|EPI_ISL_559440\|20 | ···················· | ···················· | ····················· | ···················· | ···················· | ···················· |
| J | 8176 hCoV-19/England/ALDP-49E4DA/2020\|EPI_ISL_559441\|20 | ···················· | ···················· | ····················· | ···················· | ···················· | ···················· |
| J | 8177 hCoV-19/England/ALDP-49DFCB/2020\|EPI_ISL_559442\|20 | ···················· | ···················· | ····················· | ···················· | ···················· | ···················· |
| J | 8178 hCoV-19/England/ALDP-4A47BA/2020\|EPI_ISL_559443\|20 | ···················· | ···················· | ····················· | ···················· | ···················· | ···················· |
| J | 8179 hCoV-19/England/ALDP-49F169/2020\|EPI_ISL_559444\|20 | ···················· | ···················· | ····················· | ···················· | ···················· | ···················· |
| J | 8180 hCoV-19/England/ALDP-4A4A57/2020\|EPI_ISL_559445\|20 | ···················· | ···················· | ····················· | ···················· | ···················· | ···················· |
| J | 8181 hCoV-19/England/ALDP-4ACA13/2020\|EPI_ISL_559446\|20 | ···················· | ···················· | ····················· | ···················· | ···················· | ···················· |
| J | 8182 hCoV-19/England/ALDP-4AD010/2020\|EPI_ISL_559448\|20 | ···················· | ···················· | ····················· | ···················· | ···················· | ···················· |
| J | 8183 hCoV-19/England/ALDP-49E8FC/2020\|EPI_ISL_559449\|20 | ···················· | ···················· | ····················· | ···················· | ···················· | ···················· |
| J | 8184 hCoV-19/England/ALDP-4A79DF/2020\|EPI_ISL_559450\|20 | ···················· | ···················· | ····················· | ···················· | ···················· | ···················· |
| J | 8185 hCoV-19/England/ALDP-49ED57/2020\|EPI_ISL_559451\|20 | ···················· | ···················· | ····················· | ···················· | ···················· | ···················· |
| J | 8186 hCoV-19/England/ALDP-4A5AA1/2020\|EPI_ISL_559452\|20 | ···················· | ···················· | ····················· | ···················· | ···················· | ···················· |
| J | 8187 hCoV-19/England/ALDP-4AA74B/2020\|EPI_ISL_559453\|20 | ···················· | ···················· | ····················· | ···················· | ···················· | ···················· |
| J | 8188 hCoV-19/England/ALDP-4AC873/2020\|EPI_ISL_559454\|20 | ···················· | ···················· | ····················· | ···················· | ···················· | ···················· |
| J | 8189 hCoV-19/England/ALDP-4ACE44/2020\|EPI_ISL_559455\|20 | ···················· | ···················· | ····················· | ···················· | ···················· | ···················· |
| J | 8190 hCoV-19/England/ALDP-49F530/2020\|EPI_ISL_559457\|20 | ···················· | ···················· | ····················· | ···················· | ···················· | ···················· |
| J | 8191 hCoV-19/England/ALDP-4A7526/2020\|EPI_ISL_559458\|20 | ···················· | ···················· | ····················· | ···················· | ···················· | ···················· |
| J | 8192 hCoV-19/England/ALDP-49DA42/2020\|EPI_ISL_559459\|20 | ···················· | ···················· | ····················· | ···················· | ···················· | ···················· |
| J | 8193 hCoV-19/England/MILK-99D6EF/2020\|EPI_ISL_549936\|20 | ···················· | ···················· | ····················· | ···················· | ···················· | ···················· |
| J | 8194 hCoV-19/England/MILK-99D6A3/2020\|EPI_ISL_549937\|20 | ···················· | ···················· | ····················· | ···················· | ···················· | ···················· |
| J | 8195 hCoV-19/England/MILK-99D10C/2020\|EPI_ISL_549938\|20 | ···················· | ·A·················· | ····················· | ···················· | ···················· | ···················· |
| J | 8196 hCoV-19/England/MILK-99CE33/2020\|EPI_ISL_549939\|20 | ···················· | ···················· | ····················· | ···················· | ···················· | ···················· |
| J | 8197 hCoV-19/Wales/MILK-99CDDC/2020\|EPI_ISL_549940\|2020 | ···················· | ···················· | ····················· | ···················· | ···················· | ···················· |
| J | 8198 hCoV-19/England/MILK-99CCEE/2020\|EPI_ISL_549941\|20 | ···················· | ···················· | ····················· | ···················· | ···················· | ···················· |
| J | 8199 hCoV-19/England/MILK-99D342/2020\|EPI_ISL_549942\|20 | ···················· | ···················· | ····················· | ···················· | ···················· | ···················· |
| J | 8200 hCoV-19/England/MILK-99CD81/2020\|EPI_ISL_549943\|20 | ···················· | ···················· | ····················· | ···················· | ···················· | ···················· |
| J | 8201 hCoV-19/England/MILK-99CDAF/2020\|EPI_ISL_549944\|20 | ···················· | ···················· | ····················· | ···················· | ···················· | ···················· |
| J | 8202 hCoV-19/England/MILK-99D148/2020\|EPI_ISL_549945\|20 | ···················· | ···················· | ····················· | ···················· | ···················· | ···················· |
| J | 8203 hCoV-19/England/MILK-99CC93/2020\|EPI_ISL_549947\|20 | ········T··········· | ···················· | ····················· | ···················· | ···················· | ···················· |
| J | 8204 hCoV-19/England/MILK-9A6D84/2020\|EPI_ISL_549948\|20 | ···················· | ···················· | ····················· | ···················· | ···················· | ···················· |
| J | 8205 hCoV-19/England/MILK-9A8C76/2020\|EPI_ISL_549949\|20 | ···················· | ···················· | ····················· | ···················· | ···················· | ···················· |
| J | 8206 hCoV-19/England/MILK-9A6AF6/2020\|EPI_ISL_549950\|20 | ···················· | ···················· | ····················· | ···················· | ···················· | ···················· |
| J | 8207 hCoV-19/England/MILK-9A523B/2020\|EPI_ISL_549951\|20 | ···················· | ·A·······A·········· | ····················· | ···················· | ···················· | ···················· |
| J | 8208 hCoV-19/England/MILK-9A52C2/2020\|EPI_ISL_549952\|20 | ···················· | ·A·················· | ····················· | ···················· | ···················· | ···A················ |
| J | 8209 hCoV-19/England/MILK-9A6B4E/2020\|EPI_ISL_549954\|20 | ···················· | ·A·················· | ····················· | ···················· | ···················· | ···A················ |
| J | 8210 hCoV-19/England/MILK-9A6759/2020\|EPI_ISL_549955\|20 | ···················· | ···················· | ····················· | ···················· | ···················· | ···················· |
| J | 8211 hCoV-19/Wales/MILK-9A53DE/2020\|EPI_ISL_549956\|2020 | ···················· | ···················· | ····················· | ···················· | ···················· | ···················· |
| J | 8212 hCoV-19/England/MILK-9A6BE4/2020\|EPI_ISL_549957\|20 | ···················· | ···················· | ····················· | ···················· | ···················· | ···················· |
| J | 8213 hCoV-19/England/MILK-9A55C9/2020\|EPI_ISL_549958\|20 | ···················· | ···················· | ····················· | ···················· | ···················· | ···················· |
| J | 8214 hCoV-19/England/MILK-9A557E/2020\|EPI_ISL_549959\|20 | ···················· | ···················· | ····················· | ···················· | ···················· | ···················· |
| J | 8215 hCoV-19/England/MILK-9A5611/2020\|EPI_ISL_549960\|20 | ···················· | ···················· | ····················· | ···················· | ···················· | ···················· |
| J | 8216 hCoV-19/England/MILK-9A6698/2020\|EPI_ISL_549961\|20 | ···················· | ···················· | ····················· | ···················· | ···················· | ···················· |
| J | 8217 hCoV-19/England/MILK-9A6EDC/2020\|EPI_ISL_549962\|20 | ···················· | ···················· | ····················· | ···················· | ···················· | ···················· |
| J | 8218 hCoV-19/England/MILK-9A51D4/2020\|EPI_ISL_549963\|20 | ···················· | ···················· | ····················· | ···················· | ···················· | ···················· |
| J | 8219 hCoV-19/England/MILK-9A6C1E/2020\|EPI_ISL_549964\|20 | ···················· | ···················· | ····················· | ···················· | ···················· | ···················· |
| J | 8220 hCoV-19/England/MILK-9A8371/2020\|EPI_ISL_549966\|20 | ···················· | ···················· | ····················· | ···················· | ···················· | ···················· |
| J | 8221 hCoV-19/England/MILK-9A6953/2020\|EPI_ISL_549967\|20 | ···················· | ···················· | ····················· | ···················· | ···················· | ···················· |
| J | 8222 hCoV-19/England/MILK-9A6847/2020\|EPI_ISL_549968\|20 | ···················· | ···················· | ····················· | ···················· | ···················· | ···················· |
| J | 8223 hCoV-19/England/MILK-9A505F/2020\|EPI_ISL_549969\|20 | ···················· | ···················· | ····················· | ···················· | ···················· | ···················· |
| J | 8224 hCoV-19/England/MILK-9A6856/2020\|EPI_ISL_549970\|20 | ···················· | ···················· | ····················· | ···················· | ···················· | ···················· |
| J | 8225 hCoV-19/England/MILK-9A8B6A/2020\|EPI_ISL_549971\|20 | ···················· | ···················· | ····················· | ···················· | ···················· | ···················· |
| J | 8226 hCoV-19/England/MILK-9A6926/2020\|EPI_ISL_549973\|20 | ···················· | ···················· | ····················· | ···················· | ···················· | ···················· |
| J | 8227 hCoV-19/England/MILK-9A6A05/2020\|EPI_ISL_549974\|20 | ···················· | ···················· | ····················· | ···················· | ···················· | ···················· |
| J | 8228 hCoV-19/England/MILK-9A8CD0/2020\|EPI_ISL_549975\|20 | ···················· | ···················· | ····················· | ···················· | ···················· | ···················· |
| J | 8229 hCoV-19/England/MILK-9A83AE/2020\|EPI_ISL_549977\|20 | ···················· | ···················· | ····················· | ···················· | ···················· | ···················· |
| J | 8230 hCoV-19/England/MILK-9A50E6/2020\|EPI_ISL_549978\|20 | ···················· | ···················· | ····················· | ···················· | ···················· | ···················· |
| J | 8231 hCoV-19/Wales/MILK-9A8696/2020\|EPI_ISL_549979\|2020 | ···················· | ···················· | ····················· | ···················· | ···················· | ···················· |
| J | 8232 hCoV-19/England/MILK-9A848D/2020\|EPI_ISL_549980\|20 | ···················· | ···················· | ····················· | ···················· | ···················· | ···················· |
| J | 8233 hCoV-19/England/MILK-9A54F9/2020\|EPI_ISL_549982\|20 | ···················· | ···················· | ····················· | ···················· | ···················· | ···················· |
| J | 8234 hCoV-19/England/MILK-9A51F2/2020\|EPI_ISL_549983\|20 | ···················· | ···················· | ····················· | ···················· | ···················· | ···················· |

| | | | | | | | |
|---|---|---|---|---|---|---|---|
| J | 8235 hCoV-19/England/MILK-9A6E90/2020\|EPI_ISL_549985\|20 | ···················· | ···················· | ···················· | ···················· | ······T············· | ···················· |
| J | 8236 hCoV-19/England/MILK-9A8BD3/2020\|EPI_ISL_549986\|20 | ···················· | ·A·················· | ···················· | ···················· | ···················· | ···A················ |
| J | 8237 hCoV-19/England/MILK-9A8502/2020\|EPI_ISL_549988\|20 | ···················· | ···················· | ···················· | ···················· | ···················· | ···················· |
| J | 8238 hCoV-19/England/MILK-9A86B4/2020\|EPI_ISL_549989\|20 | ···················· | ···················· | ···················· | ···················· | ···················· | ···················· |
| J | 8239 hCoV-19/England/MILK-9AA746/2020\|EPI_ISL_550920\|20 | ···················· | ···················· | ···················· | ···················· | ···················· | ···················· |
| J | 8240 hCoV-19/England/MILK-9AAAE3/2020\|EPI_ISL_550921\|20 | ···················· | ···················· | ···················· | ···················· | ···················· | ···················· |
| J | 8241 hCoV-19/England/MILK-9AAD26/2020\|EPI_ISL_550922\|20 | ···················· | ···················· | ···················· | ···················· | ···················· | ···················· |
| J | 8242 hCoV-19/England/MILK-9AA96E/2020\|EPI_ISL_550923\|20 | ···················· | ···················· | ···················· | ···················· | ···················· | ···················· |
| J | 8243 hCoV-19/England/MILK-9AA5B5/2020\|EPI_ISL_550924\|20 | ···················· | ···················· | ···················· | ···················· | ···················· | ···················· |
| J | 8244 hCoV-19/England/MILK-9AA8CB/2020\|EPI_ISL_550925\|20 | ···················· | ·A·················· | ···················· | ···················· | ···················· | ···A················ |
| J | 8245 hCoV-19/England/MILK-9A9914/2020\|EPI_ISL_550926\|20 | ···················· | ···················· | ···················· | ···················· | ···················· | ···················· |
| J | 8246 hCoV-19/England/MILK-9AA175/2020\|EPI_ISL_550927\|20 | ···················· | ······C············· | ···················· | ···················· | ···················· | ···················· |
| J | 8247 hCoV-19/England/MILK-9AA3F7/2020\|EPI_ISL_550928\|20 | ···················· | ···················· | ···················· | ···················· | ···················· | ···················· |
| J | 8248 hCoV-19/England/MILK-9AA4F4/2020\|EPI_ISL_550930\|20 | ···················· | ···················· | ···················· | ···················· | ···················· | ···················· |
| J | 8249 hCoV-19/England/MILK-9AA403/2020\|EPI_ISL_550931\|20 | ···················· | ···················· | ···················· | ···················· | ···················· | ···················· |
| J | 8250 hCoV-19/England/MILK-9AA2FA/2020\|EPI_ISL_550932\|20 | ···················· | ·A·················· | ···················· | ···················· | ···················· | ···A················ |
| J | 8251 hCoV-19/England/MILK-9AA913/2020\|EPI_ISL_550934\|20 | ···················· | ·A·················· | ···················· | ···················· | ···················· | ···················· |
| J | 8252 hCoV-19/England/MILK-9AA272/2020\|EPI_ISL_550937\|20 | ···················· | ···················· | ···················· | ···················· | ···················· | ···················· |
| J | 8253 hCoV-19/England/MILK-9AA184/2020\|EPI_ISL_550938\|20 | ···················· | ······C············· | ···················· | ···················· | ···················· | ···················· |
| J | 8254 hCoV-19/England/MILK-9A9F5E/2020\|EPI_ISL_550939\|20 | ···················· | ···················· | ···················· | ···················· | ···················· | ···················· |
| J | 8255 hCoV-19/England/MILK-9AAB77/2020\|EPI_ISL_550940\|20 | ···················· | ···················· | ···················· | ···················· | ···················· | ···················· |
| J | 8256 hCoV-19/England/MILK-9AA931/2020\|EPI_ISL_550941\|20 | ···················· | ···················· | ···················· | ···················· | ···················· | ···················· |
| J | 8257 hCoV-19/England/MILK-9AA39D/2020\|EPI_ISL_550942\|20 | ···················· | ···················· | ···················· | ···················· | ···················· | ···················· |
| J | 8258 hCoV-19/England/MILK-9AA55B/2020\|EPI_ISL_550943\|20 | ···················· | ···················· | ···················· | ···················· | ···················· | ···················· |
| J | 8259 hCoV-19/England/MILK-9AA6FE/2020\|EPI_ISL_550944\|20 | ···················· | ···················· | ···················· | ···················· | ···················· | ···················· |
| J | 8260 hCoV-19/Wales/MILK-9AA6B2/2020\|EPI_ISL_550945\|2020 | ···················· | ···················· | ···················· | ···················· | ···················· | ···················· |
| J | 8261 hCoV-19/England/MILK-9AA0C3/2020\|EPI_ISL_550946\|20 | ···················· | ···················· | ···················· | ···················· | ···················· | ···················· |
| J | 8262 hCoV-19/England/MILK-9AA5A6/2020\|EPI_ISL_550947\|20 | ···················· | ···················· | ···················· | ···················· | ···················· | ···················· |
| J | 8263 hCoV-19/England/MILK-9AA078/2020\|EPI_ISL_550948\|20 | ···················· | ···················· | ···················· | ···················· | ···················· | ···················· |
| J | 8264 hCoV-19/England/MILK-9AA0B4/2020\|EPI_ISL_550949\|20 | ···················· | ···················· | ···················· | ···················· | ···················· | ···················· |
| J | 8265 hCoV-19/England/MILK-9AADAE/2020\|EPI_ISL_550950\|20 | ···················· | ···················· | ···················· | ···················· | ···················· | ···················· |
| J | 8266 hCoV-19/England/MILK-9AABC2/2020\|EPI_ISL_550951\|20 | ···················· | ···················· | ···················· | ···················· | ···················· | ···················· |
| J | 8267 hCoV-19/England/MILK-9AAC0B/2020\|EPI_ISL_550953\|20 | ···················· | ···················· | ···················· | ···················· | ···················· | ···················· |
| J | 8268 hCoV-19/England/MILK-9AA37F/2020\|EPI_ISL_550954\|20 | ···················· | ···················· | ···················· | ···················· | ···················· | ···················· |
| J | 8269 hCoV-19/England/MILK-9AA49A/2020\|EPI_ISL_550955\|20 | ···················· | ···················· | ···················· | ···················· | ···················· | ···················· |
| J | 8270 hCoV-19/England/MILK-9AAA7A/2020\|EPI_ISL_550956\|20 | ···················· | ···················· | ···················· | ···················· | ···················· | ···················· |
| J | 8271 hCoV-19/England/MILK-9AA870/2020\|EPI_ISL_550957\|20 | ···················· | ···················· | ···················· | ···················· | ···················· | ···················· |
| J | 8272 hCoV-19/England/MILK-9AAD9F/2020\|EPI_ISL_550958\|20 | ···················· | ···················· | ···················· | ···················· | ···················· | ···················· |
| J | 8273 hCoV-19/England/MILK-9AAA98/2020\|EPI_ISL_550959\|20 | ···················· | ···················· | ···················· | ···················· | ···················· | ···················· |
| J | 8274 hCoV-19/England/MILK-9AA764/2020\|EPI_ISL_550960\|20 | ···················· | ···················· | ···················· | ···················· | ···················· | ···················· |
| J | 8275 hCoV-19/England/MILK-9AA38E/2020\|EPI_ISL_550961\|20 | ···················· | ···················· | ···················· | ···················· | ···················· | ···················· |
| J | 8276 hCoV-19/England/MILK-9AAB3B/2020\|EPI_ISL_550962\|20 | ···················· | ···················· | ···················· | ···················· | ···················· | ···················· |
| J | 8277 hCoV-19/England/MILK-9A98BD/2020\|EPI_ISL_550963\|20 | ···················· | ···················· | ···················· | ···················· | ···················· | ···················· |
| J | 8278 hCoV-19/England/MILK-9AA45E/2020\|EPI_ISL_550964\|20 | ···················· | ···················· | ···················· | ···················· | ···················· | ···················· |
| J | 8279 hCoV-19/England/MILK-9AA667/2020\|EPI_ISL_550965\|20 | ···················· | ···················· | ···················· | ···················· | ···················· | ···················· |
| J | 8280 hCoV-19/England/MILK-9AABE0/2020\|EPI_ISL_550966\|20 | ···················· | ···················· | ···················· | ···················· | ···················· | ···················· |
| J | 8281 hCoV-19/England/MILK-9AA9B9/2020\|EPI_ISL_550967\|20 | ···················· | ···················· | ···················· | ···················· | ···················· | ···················· |
| J | 8282 hCoV-19/England/MILK-9AA719/2020\|EPI_ISL_550968\|20 | ···················· | ···················· | ···················· | ···················· | ···················· | ···················· |
| J | 8283 hCoV-19/England/MILK-9AA1FD/2020\|EPI_ISL_550969\|20 | ···················· | ···················· | ···················· | ···················· | ···················· | ···················· |
| J | 8284 hCoV-19/England/MILK-9AA8AD/2020\|EPI_ISL_550980\|20 | ···················· | ·A·················· | ···················· | ···················· | ···················· | ···A················ |
| J | 8285 hCoV-19/England/MILK-992B81/2020\|EPI_ISL_552004\|20 | ···················· | ···················· | ···················· | ···················· | ···················· | ···················· |
| J | 8286 hCoV-19/England/CAMC-9915B1/2020\|EPI_ISL_552005\|20 | ···················· | ···················· | ···················· | ···················· | ···················· | ···················· |
| J | 8287 hCoV-19/England/MILK-993670/2020\|EPI_ISL_552007\|20 | ···················· | ···················· | ···················· | ···················· | ···················· | ···················· |
| J | 8288 hCoV-19/England/CAMC-991593/2020\|EPI_ISL_552008\|20 | ···················· | ···················· | ···················· | ···················· | ···················· | ···················· |
| J | 8289 hCoV-19/England/CAMC-991487/2020\|EPI_ISL_552009\|20 | ···················· | ···················· | ···················· | ···················· | ···················· | ···················· |
| J | 8290 hCoV-19/England/CAMC-99136C/2020\|EPI_ISL_552010\|20 | ···················· | ···················· | ···················· | ···················· | ···················· | ···················· |
| J | 8291 hCoV-19/England/MILK-993009/2020\|EPI_ISL_552016\|20 | ···················· | ···················· | ···················· | ···················· | ···················· | ···················· |
| J | 8292 hCoV-19/England/CAMC-9916EB/2020\|EPI_ISL_552017\|20 | ···················· | ···················· | ···················· | ···················· | ···················· | ···················· |
| J | 8293 hCoV-19/England/MILK-992FD0/2020\|EPI_ISL_552018\|20 | ···················· | ···················· | ···················· | ···················· | ···················· | ···················· |
| J | 8294 hCoV-19/England/MILK-9926AE/2020\|EPI_ISL_552019\|20 | ···················· | ···················· | ···················· | ···················· | ···················· | ···················· |
| J | 8295 hCoV-19/England/MILK-99318E/2020\|EPI_ISL_552020\|20 | ···················· | ···················· | ···················· | ···················· | ···················· | ···················· |
| J | 8296 hCoV-19/England/MILK-992662/2020\|EPI_ISL_552021\|20 | ···················· | ···················· | ···················· | ···················· | ···················· | ···················· |
| J | 8297 hCoV-19/England/MILK-993731/2020\|EPI_ISL_552023\|20 | ···················· | ···················· | ···················· | ···················· | ···················· | ···················· |
| J | 8298 hCoV-19/England/MILK-992C33/2020\|EPI_ISL_552024\|20 | ···················· | ···················· | ···················· | ···················· | ···················· | ···················· |
| J | 8299 hCoV-19/England/ALDP-90C5EE/2020\|EPI_ISL_553335\|20 | ···················· | ···················· | ···················· | ···················· | ···················· | ···················· |
| J | 8300 hCoV-19/England/ALDP-91CF92/2020\|EPI_ISL_553337\|20 | ···················· | ···················· | ···················· | ···················· | ···················· | ···················· |
| J | 8301 hCoV-19/England/ALDP-924735/2020\|EPI_ISL_553338\|20 | ···················· | ···················· | ···················· | ···················· | ···················· | ···················· |
| J | 8302 hCoV-19/England/ALDP-921E7F/2020\|EPI_ISL_553339\|20 | ···················· | ···················· | ···················· | ···················· | ···················· | ···················· |
| J | 8303 hCoV-19/England/ALDP-91F885/2020\|EPI_ISL_553340\|20 | ···················· | ···················· | ···················· | ···················· | ···················· | ···················· |
| J | 8304 hCoV-19/England/ALDP-91E82C/2020\|EPI_ISL_553341\|20 | ···················· | ···················· | ···················· | ···················· | ···················· | ···················· |
| J | 8305 hCoV-19/England/ALDP-915184/2020\|EPI_ISL_553342\|20 | ···················· | ···················· | ···················· | ···················· | ···················· | ···················· |
| J | 8306 hCoV-19/England/ALDP-922500/2020\|EPI_ISL_553343\|20 | ···················· | ···················· | ···················· | ···················· | ···················· | ···················· |
| J | 8307 hCoV-19/England/ALDP-95D26F/2020\|EPI_ISL_553346\|20 | ···················· | ···················· | ···················· | ···················· | ···················· | ···················· |
| J | 8308 hCoV-19/England/ALDP-954D67/2020\|EPI_ISL_553348\|20 | ···················· | ···················· | ···················· | ···················· | ···················· | ···················· |
| J | 8309 hCoV-19/England/ALDP-95A833/2020\|EPI_ISL_553351\|20 | ···················· | ···················· | ···················· | ···················· | ···················· | ···················· |
| J | 8310 hCoV-19/England/ALDP-954B30/2020\|EPI_ISL_553352\|20 | ···················· | ·A·················· | ···················· | ···················· | ···················· | ···················· |
| J | 8311 hCoV-19/England/ALDP-954D49/2020\|EPI_ISL_553353\|20 | ···················· | ·A·················· | ···················· | ···················· | ···················· | ···················· |
| J | 8312 hCoV-19/England/ALDP-95C8D7/2020\|EPI_ISL_553354\|20 | ···················· | ···················· | ···················· | ···················· | ···················· | ···················· |
| J | 8313 hCoV-19/England/ALDP-95CF2D/2020\|EPI_ISL_553355\|20 | ···················· | ···················· | ···················· | ···················· | ···················· | ···················· |
| J | 8314 hCoV-19/England/MILK-9584C8/2020\|EPI_ISL_554668\|20 | ···················· | ···················· | ···················· | ···················· | ···················· | ···················· |

| | | | | | | | |
|---|---|---|---|---|---|---|---|
| J | 8315 hCoV-19/England/MILK-9585A7/2020\|EPI_ISL_554670\|20 | ···················· | ···················· | ···················· | ···················· | ···················· | ···················· |
| J | 8316 hCoV-19/England/MILK-957F40/2020\|EPI_ISL_554671\|20 | ···················· | ···················· | ···················· | ···················· | ···················· | ···················· |
| J | 8317 hCoV-19/England/MILK-958792/2020\|EPI_ISL_554672\|20 | ···················· | ···················· | ···················· | ···················· | ···················· | ···················· |
| J | 8318 hCoV-19/England/MILK-957E16/2020\|EPI_ISL_554673\|20 | ···················· | ···················· | ···················· | ···················· | ···················· | ···················· |
| J | 8319 hCoV-19/England/MILK-9589C9/2020\|EPI_ISL_554674\|20 | ···················· | ···················· | ···················· | ···················· | ···················· | ···················· |
| J | 8320 hCoV-19/England/ALDP-9440AD/2020\|EPI_ISL_554677\|20 | ···················· | ·A·················· | ···················· | ···················· | ···················· | ···················· |
| J | 8321 hCoV-19/England/MILK-958255/2020\|EPI_ISL_554678\|20 | ···················· | ···················· | ···················· | ···················· | ···················· | ···················· |
| J | 8322 hCoV-19/England/MILK-957ECB/2020\|EPI_ISL_554679\|20 | ···················· | ···················· | ···················· | ···················· | ···················· | ···················· |
| J | 8323 hCoV-19/England/ALDP-94430E/2020\|EPI_ISL_554680\|20 | ···················· | ···················· | ···················· | ···················· | ···················· | ···················· |
| J | 8324 hCoV-19/England/ALDP-944210/2020\|EPI_ISL_554681\|20 | ···················· | ···················· | ···················· | ···················· | ···················· | ···················· |
| J | 8325 hCoV-19/England/MILK-958273/2020\|EPI_ISL_554682\|20 | ···················· | ···················· | ···················· | ···················· | ···················· | ···················· |
| J | 8326 hCoV-19/England/MILK-957C76/2020\|EPI_ISL_554683\|20 | ···················· | ···················· | ···················· | ···················· | ···················· | ···················· |
| J | 8327 hCoV-19/England/MILK-9587ED/2020\|EPI_ISL_554684\|20 | ···················· | ···················· | ···················· | ···················· | ···················· | ···················· |
| J | 8328 hCoV-19/England/MILK-957CC1/2020\|EPI_ISL_554685\|20 | ···················· | ···················· | ···················· | ···················· | ···················· | ···················· |
| J | 8329 hCoV-19/England/ALDP-94469C/2020\|EPI_ISL_554686\|20 | ···················· | ·A·················· | ···················· | ···················· | ···················· | ···················· |
| J | 8330 hCoV-19/England/MILK-9581EF/2020\|EPI_ISL_554687\|20 | ···················· | ···················· | ···················· | ···················· | ···················· | ···················· |
| J | 8331 hCoV-19/England/ALDP-94C807/2020\|EPI_ISL_555900\|20 | ···················· | ···················· | ···················· | ···················· | ···················· | ···················· |
| J | 8332 hCoV-19/England/ALDP-94C61C/2020\|EPI_ISL_555901\|20 | ···················· | ···················· | ···················· | ···················· | ···················· | ···················· |
| J | 8333 hCoV-19/England/ALDP-94CB95/2020\|EPI_ISL_555902\|20 | ···················· | ·A·················· | ···················· | ···················· | ···················· | ···················· |
| J | 8334 hCoV-19/England/ALDP-94C5D3/2020\|EPI_ISL_555903\|20 | ···················· | ···················· | ···················· | ···················· | ···················· | ···················· |
| J | 8335 hCoV-19/England/ALDP-94CAB6/2020\|EPI_ISL_555904\|20 | ···················· | ···················· | ···················· | ···················· | ···················· | ···················· |
| J | 8336 hCoV-19/England/ALDP-94C922/2020\|EPI_ISL_555907\|20 | ···················· | ···················· | ···················· | ···················· | ···················· | ···················· |
| J | 8337 hCoV-19/England/ALDP-94CE32/2020\|EPI_ISL_555908\|20 | ···················· | ···················· | ···················· | ···················· | ···················· | ···················· |
| J | 8338 hCoV-19/England/ALDP-94CC29/2020\|EPI_ISL_555909\|20 | ···················· | ···················· | ···················· | ···················· | ···················· | ···················· |
| J | 8339 hCoV-19/England/CAMC-949BD4/2020\|EPI_ISL_555910\|20 | ···················· | ·A·················· | ···················· | ···················· | ···················· | ··A················· |
| J | 8340 hCoV-19/England/ALDP-94C49A/2020\|EPI_ISL_555911\|20 | ···················· | ···················· | ···················· | ···················· | ···················· | ···················· |
| J | 8341 hCoV-19/England/ALDP-94D183/2020\|EPI_ISL_555912\|20 | ···················· | ···················· | ···················· | ···················· | ···················· | ···················· |
| J | 8342 hCoV-19/England/ALDP-94CD35/2020\|EPI_ISL_555913\|20 | ···················· | ···················· | ···················· | ···················· | ···················· | ···················· |
| J | 8343 hCoV-19/England/ALDP-94CB77/2020\|EPI_ISL_555914\|20 | ···················· | ···················· | ···················· | ···················· | ···················· | ···················· |
| J | 8344 hCoV-19/England/ALDP-94C5A6/2020\|EPI_ISL_555915\|20 | ···················· | ···················· | ···················· | ···················· | ···················· | ···················· |
| J | 8345 hCoV-19/England/ALDP-94CBE0/2020\|EPI_ISL_555917\|20 | ···················· | ···················· | ···················· | ···················· | ···················· | ···················· |
| J | 8346 hCoV-19/England/ALDP-94C8CB/2020\|EPI_ISL_555918\|20 | ···················· | ···················· | ···················· | ···················· | ···················· | ···················· |
| J | 8347 hCoV-19/England/ALDP-94C62B/2020\|EPI_ISL_555919\|20 | ···················· | ···················· | ···················· | ···················· | ···················· | ···················· |
| J | 8348 hCoV-19/England/ALDP-94C46D/2020\|EPI_ISL_555920\|20 | ···················· | ···················· | ···················· | ···················· | ···················· | ···················· |
| J | 8349 hCoV-19/England/ALDP-94CC1A/2020\|EPI_ISL_555921\|20 | ···················· | ···················· | ···················· | ···················· | ···················· | ···················· |
| J | 8350 hCoV-19/England/ALDP-94C5B5/2020\|EPI_ISL_555923\|20 | ···················· | ···················· | ···················· | ···················· | ···················· | ···················· |
| J | 8351 hCoV-19/England/ALDP-94F637/2020\|EPI_ISL_555924\|20 | ···················· | ···················· | ···················· | ···················· | ···················· | ···················· |
| J | 8352 hCoV-19/England/ALDP-94C8BC/2020\|EPI_ISL_555925\|20 | ···················· | ···················· | ···················· | ···················· | ···················· | ···················· |
| J | 8353 hCoV-19/England/ALDP-94CBB3/2020\|EPI_ISL_555926\|20 | ···················· | ···················· | ···················· | ···················· | ···················· | ···················· |
| J | 8354 hCoV-19/England/ALDP-94D129/2020\|EPI_ISL_555927\|20 | ···················· | ···················· | ···················· | ···················· | ···················· | ···················· |
| J | 8355 hCoV-19/Wales/ALDP-94C746/2020\|EPI_ISL_555928\|2020 | ···················· | ···················· | ···················· | ···················· | ···················· | ···················· |
| J | 8356 hCoV-19/England/ALDP-94C782/2020\|EPI_ISL_555930\|20 | ···················· | ···················· | ···················· | ···················· | ···················· | ···················· |
| J | 8357 hCoV-19/England/ALDP-94F479/2020\|EPI_ISL_555998\|20 | ···················· | ···················· | ···················· | ···················· | ···················· | ···················· |
| J | 8358 hCoV-19/England/ALDP-94C7BF/2020\|EPI_ISL_555999\|20 | ···················· | ···················· | ···················· | ···················· | ···················· | ···················· |
| J | 8359 hCoV-19/England/ALDP-68DEFC/2020\|EPI_ISL_558130\|20 | ···················· | ···················· | ···················· | ···················· | ···················· | ···················· |
| J | 8360 hCoV-19/England/ALDP-658C78/2020\|EPI_ISL_558132\|20 | ···················· | ···················· | ···················· | ···················· | ···················· | ···················· |
| J | 8361 hCoV-19/England/ALDP-66DEE3/2020\|EPI_ISL_558133\|20 | ···················· | ···················· | ···················· | ···················· | ···················· | ···················· |
| J | 8362 hCoV-19/England/ALDP-68C938/2020\|EPI_ISL_558134\|20 | ···················· | ···················· | ···················· | ···················· | ···················· | ···················· |
| J | 8363 hCoV-19/England/ALDP-643F3B/2020\|EPI_ISL_558136\|20 | ···················· | ···················· | ···················· | ···················· | ···················· | ···················· |
| J | 8364 hCoV-19/England/ALDP-66B137/2020\|EPI_ISL_558137\|20 | ···················· | ···················· | ···················· | ···················· | ···················· | ···················· |
| J | 8365 hCoV-19/England/ALDP-644D9A/2020\|EPI_ISL_558139\|20 | ···················· | ···················· | ···················· | ···················· | ···················· | ···················· |
| J | 8366 hCoV-19/England/ALDP-652F57/2020\|EPI_ISL_558140\|20 | ···················· | ···················· | ···················· | ···················· | ···················· | ···················· |
| J | 8367 hCoV-19/England/ALDP-6926ED/2020\|EPI_ISL_558142\|20 | ···················· | ···················· | ···················· | ···················· | ···················· | ···················· |
| J | 8368 hCoV-19/England/ALDP-65980A/2020\|EPI_ISL_558144\|20 | ···················· | ···················· | ···················· | ···················· | ···················· | ···················· |
| J | 8369 hCoV-19/England/ALDP-68E4BD/2020\|EPI_ISL_558145\|20 | ···················· | ···················· | ···················· | ···················· | ···················· | ···················· |
| J | 8370 hCoV-19/England/ALDP-66E6AE/2020\|EPI_ISL_558146\|20 | ···················· | ···················· | ···················· | ···················· | ···················· | ···················· |
| J | 8371 hCoV-19/England/ALDP-643F1D/2020\|EPI_ISL_558148\|20 | ···················· | ···················· | ···················· | ···················· | ···················· | ···················· |
| J | 8372 hCoV-19/England/ALDP-66E222/2020\|EPI_ISL_558151\|20 | ···················· | ···················· | ···················· | ···················· | ···················· | ···················· |
| J | 8373 hCoV-19/England/ALDP-6532E4/2020\|EPI_ISL_558153\|20 | ···················· | ···················· | ···················· | ···················· | ···················· | ···················· |
| J | 8374 hCoV-19/England/ALDP-66EE97/2020\|EPI_ISL_558154\|20 | ···················· | ···················· | ···················· | ···················· | ···················· | ···················· |
| J | 8375 hCoV-19/England/MILK-654007/2020\|EPI_ISL_558155\|20 | ···················· | ·A·················· | ···················· | ···················· | ···················· | ···················· |
| J | 8376 hCoV-19/England/ALDP-68C015/2020\|EPI_ISL_558156\|20 | ···················· | ···················· | ···················· | ···················· | ···················· | ···················· |
| J | 8377 hCoV-19/England/ALDP-647CE5/2020\|EPI_ISL_558158\|20 | ···················· | ···················· | ···················· | ···················· | ···················· | ···················· |
| J | 8378 hCoV-19/England/ALDP-6495E8/2020\|EPI_ISL_558160\|20 | ···················· | ···················· | ···················· | ···················· | ···················· | ···················· |
| J | 8379 hCoV-19/England/ALDP-69063A/2020\|EPI_ISL_558164\|20 | ···················· | ···················· | ···················· | ···················· | ···················· | ···················· |
| J | 8380 hCoV-19/England/ALDP-6749CA/2020\|EPI_ISL_558166\|20 | ···················· | ···················· | ···················· | ···················· | ···················· | ···················· |
| J | 8381 hCoV-19/England/ALDP-658829/2020\|EPI_ISL_558168\|20 | ···················· | ···················· | ···················· | ···················· | ···················· | ···················· |
| J | 8382 hCoV-19/England/ALDP-658504/2020\|EPI_ISL_558169\|20 | ···················· | ···················· | ···················· | ···················· | ···················· | ···················· |
| J | 8383 hCoV-19/England/ALDP-6517D8/2020\|EPI_ISL_558170\|20 | ···················· | ···················· | ···················· | ···················· | ···················· | ···················· |
| J | 8384 hCoV-19/England/ALDP-6765C4/2020\|EPI_ISL_558171\|20 | ···················· | ···················· | ···················· | ···················· | ···················· | ···················· |
| J | 8385 hCoV-19/England/ALDP-646BE9/2020\|EPI_ISL_558172\|20 | ···················· | ···················· | ···················· | ···················· | ···················· | ···················· |
| J | 8386 hCoV-19/England/ALDP-644F1C/2020\|EPI_ISL_558173\|20 | ···················· | ···················· | ···················· | ···················· | ···················· | ···················· |
| J | 8387 hCoV-19/Wales/ALDP-643F68/2020\|EPI_ISL_558174\|2020 | ···················· | ···················· | ···················· | ···················· | ···················· | ···················· |
| J | 8388 hCoV-19/England/ALDP-68B91B/2020\|EPI_ISL_558176\|20 | ···················· | ···················· | ···················· | ···················· | ···················· | ···················· |
| J | 8389 hCoV-19/England/ALDP-6507D9/2020\|EPI_ISL_558180\|20 | ···················· | ···················· | ···················· | ···················· | ···················· | ···················· |
| J | 8390 hCoV-19/England/ALDP-65C209/2020\|EPI_ISL_558181\|20 | ···················· | ···················· | ···················· | ···················· | ···················· | ···················· |
| J | 8391 hCoV-19/England/ALDP-651BAF/2020\|EPI_ISL_558182\|20 | ···················· | ···················· | ···················· | ···················· | ···················· | ···················· |
| J | 8392 hCoV-19/England/ALDP-68CF45/2020\|EPI_ISL_558183\|20 | ···················· | ···················· | ···················· | ···················· | ···················· | ···················· |
| J | 8393 hCoV-19/England/ALDP-68BD3D/2020\|EPI_ISL_558184\|20 | ···················· | ···················· | ···················· | ···················· | ···················· | ···················· |
| J | 8394 hCoV-19/England/ALDP-671A42/2020\|EPI_ISL_558186\|20 | ···················· | ···················· | ···················· | ···················· | ···················· | ···················· |

| | | | | | | | | |
|---|---|---|---|---|---|---|---|---|
| J | 8395 | hCoV-19/England/ALDP-692BFD/2020\|EPI_ISL_558187\|20 | ···················· | ···················· | ····················· | ···················· | ···················· | ···················· |
| J | 8396 | hCoV-19/England/ALDP-68D8B2/2020\|EPI_ISL_558189\|20 | ···················· | ···················· | ····················· | ···················· | ···················· | ···················· |
| J | 8397 | hCoV-19/England/ALDP-66EA48/2020\|EPI_ISL_558190\|20 | ···················· | ···················· | ····················· | ···················· | ···················· | ···················· |
| J | 8398 | hCoV-19/England/ALDP-65BF30/2020\|EPI_ISL_558191\|20 | ···················· | ···················· | ····················· | ···················· | ···················· | ···················· |
| J | 8399 | hCoV-19/England/ALDP-64AB30/2020\|EPI_ISL_558192\|20 | ···················· | ···················· | ····················· | ···················· | ···················· | ···················· |
| J | 8400 | hCoV-19/England/ALDP-655E39/2020\|EPI_ISL_558193\|20 | ···················· | ···················· | ····················· | ···················· | ···················· | ···················· |
| J | 8401 | hCoV-19/England/MILK-6754AA/2020\|EPI_ISL_558195\|20 | ···················· | ···················· | ····················· | ···················· | ···················· | ···················· |
| J | 8402 | hCoV-19/England/ALDP-658A8D/2020\|EPI_ISL_558196\|20 | ···················· | ···················· | ····················· | ···················· | ···················· | ···················· |
| J | 8403 | hCoV-19/England/ALDP-650B64/2020\|EPI_ISL_558197\|20 | ···················· | ···················· | ····················· | ···················· | ···················· | ···················· |
| J | 8404 | hCoV-19/England/ALDP-690FA8/2020\|EPI_ISL_558198\|20 | ···················· | ···················· | ····················· | ···················· | ···················· | ···················· |
| J | 8405 | hCoV-19/England/ALDP-672355/2020\|EPI_ISL_558199\|20 | ···················· | ···················· | ····················· | ···················· | ···················· | ···················· |
| J | 8406 | hCoV-19/England/ALDP-4AD669/2020\|EPI_ISL_559308\|20 | ···················· | ···················· | ····················· | ···················· | ···················· | ···················· |
| J | 8407 | hCoV-19/England/ALDP-49B033/2020\|EPI_ISL_559309\|20 | ···················· | ···················· | ····················· | ···················· | ···················· | ···················· |
| J | 8408 | hCoV-19/England/ALDP-4AEA99/2020\|EPI_ISL_559310\|20 | ···················· | ···················· | ····················· | ···················· | ···················· | ···················· |
| J | 8409 | hCoV-19/England/ALDP-496AD0/2020\|EPI_ISL_559311\|20 | ···················· | ···················· | ····················· | ···················· | ···················· | ···················· |
| J | 8410 | hCoV-19/England/MILK-4A0095/2020\|EPI_ISL_559312\|20 | ···················· | ···················· | ····················· | ···················· | ···················· | ···················· |
| J | 8411 | hCoV-19/England/ALDP-49687C/2020\|EPI_ISL_559313\|20 | ···················· | ···················· | ····················· | ···················· | ···················· | ···················· |
| J | 8412 | hCoV-19/England/MILK-4A0AB5/2020\|EPI_ISL_559314\|20 | ···················· | ···················· | ····················· | ···················· | ···················· | ···················· |
| J | 8413 | hCoV-19/England/ALDP-497152/2020\|EPI_ISL_559315\|20 | ···················· | ···················· | ····················· | ···················· | ···················· | ···················· |
| J | 8414 | hCoV-19/England/ALDP-49A984/2020\|EPI_ISL_559316\|20 | ···················· | ···················· | ····················· | ···················· | ···················· | ···················· |
| J | 8415 | hCoV-19/England/ALDP-496706/2020\|EPI_ISL_559317\|20 | ···················· | ···················· | ····················· | ···················· | ···················· | ···················· |
| J | 8416 | hCoV-19/England/ALDP-4ABC69/2020\|EPI_ISL_559460\|20 | ···················· | ···················· | ····················· | ···················· | ···················· | ···················· |
| J | 8417 | hCoV-19/England/ALDP-49C7D3/2020\|EPI_ISL_559462\|20 | ···················· | ···················· | ····················· | ···················· | ···················· | ···················· |
| J | 8418 | hCoV-19/England/ALDP-4A723E/2020\|EPI_ISL_559463\|20 | ···················· | ···················· | ····················· | ···················· | ···················· | ···················· |
| J | 8419 | hCoV-19/England/ALDP-4A726B/2020\|EPI_ISL_559466\|20 | ···················· | ···················· | ····················· | ···················· | ···················· | ···················· |
| J | 8420 | hCoV-19/England/ALDP-4A4B18/2020\|EPI_ISL_559467\|20 | ···················· | ···················· | ····················· | ···················· | ···················· | ···················· |
| J | 8421 | hCoV-19/England/ALDP-4A92C3/2020\|EPI_ISL_559468\|20 | ···················· | ···················· | ····················· | ···················· | ···················· | ···················· |
| J | 8422 | hCoV-19/England/ALDP-4A4BDC/2020\|EPI_ISL_559469\|20 | ···················· | ···················· | ····················· | ···················· | ···················· | ···················· |
| J | 8423 | hCoV-19/England/ALDP-4A4204/2020\|EPI_ISL_559470\|20 | ···················· | ···················· | ····················· | ···················· | ···················· | ···················· |
| J | 8424 | hCoV-19/England/ALDP-49CD86/2020\|EPI_ISL_559471\|20 | ···················· | ···················· | ····················· | ···················· | ···················· | ···················· |
| J | 8425 | hCoV-19/England/ALDP-4A931B/2020\|EPI_ISL_559472\|20 | ···················· | ···················· | ····················· | ···················· | ···················· | ···················· |
| J | 8426 | hCoV-19/England/ALDP-4A8428/2020\|EPI_ISL_559473\|20 | ···················· | ···················· | ····················· | ···················· | ···················· | ···················· |
| J | 8427 | hCoV-19/England/ALDP-4A94AF/2020\|EPI_ISL_559474\|20 | ···················· | ···················· | ····················· | ···················· | ···················· | ···················· |
| J | 8428 | hCoV-19/England/ALDP-4A4574/2020\|EPI_ISL_559475\|20 | ···················· | ···················· | ····················· | ···················· | ···················· | ···················· |
| J | 8429 | hCoV-19/England/ALDP-4A7C4F/2020\|EPI_ISL_559476\|20 | ···················· | ···················· | ····················· | ···················· | ···················· | ···················· |
| J | 8430 | hCoV-19/England/ALDP-4AD7C0/2020\|EPI_ISL_559477\|20 | ···················· | ···················· | ····················· | ···················· | ···················· | ···················· |
| J | 8431 | hCoV-19/England/ALDP-4A485D/2020\|EPI_ISL_559478\|20 | ···················· | ···················· | ····················· | ···················· | ···················· | ···················· |
| J | 8432 | hCoV-19/England/ALDP-4A4E88/2020\|EPI_ISL_559479\|20 | ···················· | ···················· | ····················· | ···················· | ···················· | ···················· |
| J | 8433 | hCoV-19/England/ALDP-4A6C9B/2020\|EPI_ISL_559480\|20 | ···················· | ···················· | ····················· | ···················· | ···················· | ···················· |
| J | 8434 | hCoV-19/England/ALDP-49D839/2020\|EPI_ISL_559483\|20 | ···················· | ···················· | ····················· | ···················· | ···················· | ···················· |
| J | 8435 | hCoV-19/England/ALDP-4A6132/2020\|EPI_ISL_559484\|20 | ···················· | ···················· | ····················· | ···················· | ···················· | ···················· |
| J | 8436 | hCoV-19/England/ALDP-4A63C3/2020\|EPI_ISL_559485\|20 | ···················· | ···················· | ····················· | ···················· | ···················· | ···················· |
| J | 8437 | hCoV-19/England/ALDP-49EE54/2020\|EPI_ISL_559486\|20 | ···················· | ···················· | ····················· | ···················· | ···················· | ···················· |
| J | 8438 | hCoV-19/England/ALDP-4ADC76/2020\|EPI_ISL_559487\|20 | ···················· | ···················· | ····················· | ···················· | ···················· | ···················· |
| J | 8439 | hCoV-19/England/ALDP-4A86E6/2020\|EPI_ISL_559488\|20 | ···················· | ···················· | ····················· | ···················· | ···················· | ···················· |
| J | 8440 | hCoV-19/England/ALDP-49E9CC/2020\|EPI_ISL_559489\|20 | ···················· | ···················· | ····················· | ···················· | ···················· | ···················· |
| J | 8441 | hCoV-19/England/ALDP-4A70BC/2020\|EPI_ISL_559491\|20 | ···················· | ···················· | ····················· | ···················· | ···················· | ···················· |
| J | 8442 | hCoV-19/England/ALDP-49E07C/2020\|EPI_ISL_559492\|20 | ···················· | ···················· | ····················· | ···················· | ···················· | ···················· |
| J | 8443 | hCoV-19/England/ALDP-4A8367/2020\|EPI_ISL_559493\|20 | ···················· | ···················· | ····················· | ···················· | ···················· | ···················· |
| J | 8444 | hCoV-19/England/ALDP-4A9E74/2020\|EPI_ISL_559495\|20 | ···················· | ···················· | ····················· | ···················· | ···················· | ···················· |
| J | 8445 | hCoV-19/England/ALDP-4A6044/2020\|EPI_ISL_559496\|20 | ···················· | ···················· | ····················· | ···················· | ···················· | ···················· |
| J | 8446 | hCoV-19/England/ALDP-4A92F0/2020\|EPI_ISL_559497\|20 | ···················· | ···················· | ····················· | ···················· | ···················· | ···················· |
| J | 8447 | hCoV-19/England/ALDP-4A50CD/2020\|EPI_ISL_559498\|20 | ···················· | ···················· | ····················· | ···················· | ···················· | ···················· |
| J | 8448 | hCoV-19/England/ALDP-4A5DA8/2020\|EPI_ISL_559499\|20 | ···················· | ···················· | ····················· | ···················· | ···················· | ···················· |
| J | 8449 | hCoV-19/England/MILK-99D649/2020\|EPI_ISL_549900\|20 | ···················· | ···················· | ····················· | ···················· | ···················· | ···················· |
| J | 8450 | hCoV-19/England/MILK-99CE60/2020\|EPI_ISL_549901\|20 | ···················· | ···················· | ····················· | ···················· | ···················· | ···················· |
| J | 8451 | hCoV-19/England/MILK-99D12A/2020\|EPI_ISL_549903\|20 | ···················· | ···················· | ····················· | ···················· | ···················· | ···················· |
| J | 8452 | hCoV-19/England/MILK-99D5F1/2020\|EPI_ISL_549904\|20 | ···················· | ···················· | ····················· | ···················· | ······T············· | ···················· |
| J | 8453 | hCoV-19/England/MILK-99CE42/2020\|EPI_ISL_549905\|20 | ···················· | ···················· | ····················· | ···················· | ···················· | ···················· |
| J | 8454 | hCoV-19/England/MILK-99D4D6/2020\|EPI_ISL_549906\|20 | ···················· | ···················· | ····················· | ···················· | ···················· | ···················· |
| J | 8455 | hCoV-19/England/MILK-99D61C/2020\|EPI_ISL_549907\|20 | ···················· | ···················· | ····················· | ···················· | ···················· | ···················· |
| J | 8456 | hCoV-19/England/MILK-99CF03/2020\|EPI_ISL_549909\|20 | ···················· | ···················· | ····················· | ···················· | ···················· | ···················· |
| J | 8457 | hCoV-19/England/MILK-99CD27/2020\|EPI_ISL_549910\|20 | ···················· | ···················· | ····················· | ···················· | ···················· | ···················· |
| J | 8458 | hCoV-19/England/MILK-99CBF0/2020\|EPI_ISL_549911\|20 | ···················· | ···················· | ····················· | ···················· | ···················· | ···················· |
| J | 8459 | hCoV-19/England/MILK-99CDBE/2020\|EPI_ISL_549912\|20 | ···················· | ···················· | ····················· | ···················· | ···················· | ···················· |
| J | 8460 | hCoV-19/England/MILK-99D755/2020\|EPI_ISL_549913\|20 | ···················· | ···················· | ····················· | ···················· | ···················· | ···················· |
| J | 8461 | hCoV-19/England/MILK-99CBB4/2020\|EPI_ISL_549914\|20 | ···················· | ···················· | ····················· | ···················· | ······T············· | ···················· |
| J | 8462 | hCoV-19/England/MILK-99CDCD/2020\|EPI_ISL_549915\|20 | ···················· | ···················· | ····················· | ···················· | ···················· | ···················· |
| J | 8463 | hCoV-19/England/MILK-99D0A5/2020\|EPI_ISL_549916\|20 | ···················· | ···················· | ····················· | ···················· | ···················· | ···················· |
| J | 8464 | hCoV-19/England/MILK-99D719/2020\|EPI_ISL_549917\|20 | ···················· | ···················· | ····················· | ···················· | ···················· | ···············A···· |
| J | 8465 | hCoV-19/England/MILK-99CECA/2020\|EPI_ISL_549918\|20 | ···················· | ···················· | ····················· | ···················· | ···················· | ···················· |
| J | 8466 | hCoV-19/England/MILK-99D60D/2020\|EPI_ISL_549921\|20 | ···················· | ···················· | ····················· | ···················· | ···················· | ···················· |
| J | 8467 | hCoV-19/England/MILK-99CC0C/2020\|EPI_ISL_549922\|20 | ···················· | ···················· | ····················· | ···················· | ···················· | ···················· |
| J | 8468 | hCoV-19/England/MILK-99D05A/2020\|EPI_ISL_549923\|20 | ···················· | ···················· | ····················· | ···················· | ···················· | ···················· |
| J | 8469 | hCoV-19/England/MILK-99CF21/2020\|EPI_ISL_549924\|20 | ···················· | ·A·················· | ····················· | ···················· | ···················· | ··A················· |
| J | 8470 | hCoV-19/England/MILK-99CF30/2020\|EPI_ISL_549925\|20 | ···················· | ···················· | ····················· | ···················· | ···················· | ···················· |
| J | 8471 | hCoV-19/England/MILK-99D5D3/2020\|EPI_ISL_549928\|20 | ···················· | ···················· | ····················· | ···················· | ···················· | ···················· |
| J | 8472 | hCoV-19/England/MILK-99D5C4/2020\|EPI_ISL_549930\|20 | ···················· | ···················· | ····················· | ···················· | ···················· | ···················· |
| J | 8473 | hCoV-19/England/MILK-99D746/2020\|EPI_ISL_549931\|20 | ···················· | ·A·················· | ····················· | ···················· | ···················· | ···················· |
| J | 8474 | hCoV-19/England/MILK-99D667/2020\|EPI_ISL_549932\|20 | ···················· | ···················· | ····················· | ···················· | ···················· | ···················· |

J 8475 hCoV-19/England/MILK-99D47C/2020|EPI_ISL_549933|20 ···················· ···················· ····················· ···················· ···················· ····················
J 8476 hCoV-19/England/MILK-99CC66/2020|EPI_ISL_549934|20 ···················· ···················· ····················· ···················· ···················· ····················
J 8477 hCoV-19/England/MILK-99D0C3/2020|EPI_ISL_549935|20 ···················· ···················· ····················· ···················· ···················· ····················
J 8478 hCoV-19/England/MILK-9A571E/2020|EPI_ISL_549990|20 ···················· ···················· ····················· ···················· ···················· ····················
J 8479 hCoV-19/England/MILK-9A54BD/2020|EPI_ISL_549991|20 ···················· ···················· ····················· ···················· ···················· ····················
J 8480 hCoV-19/England/MILK-9A52E0/2020|EPI_ISL_549992|20 ···················· ···················· ····················· ···················· ···················· ····················
J 8481 hCoV-19/England/MILK-9A6C5A/2020|EPI_ISL_549993|20 ···················· ···················· ····················· ···················· ···················· ····················
J 8482 hCoV-19/England/MILK-9A6C87/2020|EPI_ISL_549994|20 ········T··········· ···················· ····················· ···················· ···················· ····················
J 8483 hCoV-19/England/MILK-9A6D75/2020|EPI_ISL_549996|20 ···················· ···················· ····················· ···················· ···················· ····················
J 8484 hCoV-19/England/MILK-9A8D64/2020|EPI_ISL_549998|20 ···················· ···················· ····················· ···················· ···················· ····················
J 8485 hCoV-19/England/MILK-9A6E72/2020|EPI_ISL_549999|20 ···················· ···················· ····················· ···················· ···················· ····················
J 8486 hCoV-19/England/MILK-9AA5E2/2020|EPI_ISL_550900|20 ···················· ···················· ····················· ···················· ···················· ····················
J 8487 hCoV-19/Wales/MILK-9AADCC/2020|EPI_ISL_550901|2020 ···················· ···················· ····················· ···················· ···················· ····················
J 8488 hCoV-19/England/MILK-9AAC29/2020|EPI_ISL_550902|20 ···················· ···················· ····················· ···················· ···················· ····················
J 8489 hCoV-19/England/MILK-9AA00F/2020|EPI_ISL_550903|20 ···················· ···················· ····················· ···················· ···················· ····················
J 8490 hCoV-19/England/MILK-9AA3E8/2020|EPI_ISL_550904|20 ···················· ···················· ····················· ···················· ···················· ····················
J 8491 hCoV-19/England/MILK-9A9880/2020|EPI_ISL_550905|20 ···················· ···················· ····················· ···················· ···················· ····················
J 8492 hCoV-19/England/MILK-9A9F6D/2020|EPI_ISL_550906|20 ···················· ···················· ····················· ···················· ···················· ····················
J 8493 hCoV-19/England/MILK-9AAAB6/2020|EPI_ISL_550907|20 ···················· ···················· ····················· ···················· ···················· ····················
J 8494 hCoV-19/England/MILK-9AAA6B/2020|EPI_ISL_550908|20 ···················· ···················· ····················· ···················· ···················· ····················
J 8495 hCoV-19/England/MILK-9AABFF/2020|EPI_ISL_550916|20 ···················· ···················· ····················· ···················· ···················· ····················
J 8496 hCoV-19/England/MILK-9A9FD6/2020|EPI_ISL_550917|20 ···················· ···················· ····················· ···················· ···················· ····················
J 8497 hCoV-19/England/MILK-9AAD71/2020|EPI_ISL_550971|20 ···················· ···················· ····················· ···················· ···················· ····················
J 8498 hCoV-19/England/MILK-9AA62B/2020|EPI_ISL_550972|20 ···················· ···················· ····················· ···················· ···················· ····················
J 8499 hCoV-19/England/MILK-9AA9AA/2020|EPI_ISL_550973|20 ···················· ···················· ····················· ···················· ······T············· ····················
J 8500 hCoV-19/England/MILK-9A9835/2020|EPI_ISL_550974|20 ···················· ···················· ····················· ···················· ···················· ····················
J 8501 hCoV-19/England/MILK-9AA9D7/2020|EPI_ISL_550975|20 ···················· ···················· ····················· ···················· ···················· ····················
J 8502 hCoV-19/England/MILK-9AA44F/2020|EPI_ISL_550976|20 ···················· ·A·······A·········· ····················· ···················· ···················· ····················
J 8503 hCoV-19/England/MILK-9AA9E6/2020|EPI_ISL_550977|20 ···················· ···················· ····················· ···················· ···················· ····················
J 8504 hCoV-19/Wales/MILK-9AA1C0/2020|EPI_ISL_550978|2020 ···················· ···················· ····················· ···················· ···················· ····················
J 8505 hCoV-19/England/MILK-9AADF9/2020|EPI_ISL_550979|20 ···················· ···················· ····················· ···················· ···················· ····················
J 8506 hCoV-19/Wales/MILK-9AA1EE/2020|EPI_ISL_550981|2020 ···················· ···················· ····················· ···················· ···················· ····················
J 8507 hCoV-19/England/MILK-9AA816/2020|EPI_ISL_550982|20 ···················· ···················· ····················· ···················· ···················· ····················
J 8508 hCoV-19/England/MILK-9AA4B8/2020|EPI_ISL_550984|20 ···················· ···················· ····················· ···················· ······T············· ····················
J 8509 hCoV-19/England/MILK-9AA3BB/2020|EPI_ISL_550986|20 ···················· ···················· ····················· ···················· ···················· ····················
J 8510 hCoV-19/Wales/MILK-9AAD62/2020|EPI_ISL_550987|2020 ···················· ···················· ····················· ···················· ···················· ····················
J 8511 hCoV-19/England/MILK-9AA139/2020|EPI_ISL_550988|20 ···················· ···················· ····················· ···················· ···················· ····················
J 8512 hCoV-19/England/MILK-9AADEA/2020|EPI_ISL_550989|20 ···················· ·A·················· ····················· ···················· ···················· ···A················
J 8513 hCoV-19/England/MILK-9AA60D/2020|EPI_ISL_550990|20 ···················· ···················· ····················· ···················· ···················· ····················
J 8514 hCoV-19/England/MILK-9AA70A/2020|EPI_ISL_550991|20 ···················· ···················· ····················· ···················· ···················· ····················
J 8515 hCoV-19/England/MILK-9AA6EF/2020|EPI_ISL_550992|20 ···················· ···················· ····················· ···················· ···················· ····················
J 8516 hCoV-19/England/MILK-9A8EDA/2020|EPI_ISL_550993|20 ···················· ···················· ····················· ···················· ···················· ····················
J 8517 hCoV-19/England/MILK-9A8FC8/2020|EPI_ISL_550994|20 ···················· ···················· ····················· ···················· ···················· ····················
J 8518 hCoV-19/England/MILK-9A9B87/2020|EPI_ISL_550995|20 ···················· ·A·················· ····················· ···················· ···················· ····················
J 8519 hCoV-19/England/MILK-9A9A8A/2020|EPI_ISL_550996|20 ···················· ···················· ····················· ···················· ···················· ····················
J 8520 hCoV-19/England/MILK-9A9264/2020|EPI_ISL_550997|20 ···················· ···················· ····················· ···················· ···················· ····················
J 8521 hCoV-19/England/MILK-9A971A/2020|EPI_ISL_550998|20 ···················· ···················· ····················· ···················· ···················· ····················
J 8522 hCoV-19/England/MILK-9A9EF7/2020|EPI_ISL_550999|20 ···················· ···················· ····················· ···················· ···················· ····················
J 8523 hCoV-19/England/MILK-970943/2020|EPI_ISL_553115|20 ···················· ···················· ····················· ···················· ···················· ····················
J 8524 hCoV-19/England/MILK-970A8C/2020|EPI_ISL_553116|20 ···················· ···················· ····················· ···················· ···················· ····················
J 8525 hCoV-19/England/MILK-971599/2020|EPI_ISL_553117|20 ···················· ·A·················· ····················· ···················· ···················· ····················
J 8526 hCoV-19/England/MILK-9711D1/2020|EPI_ISL_553118|20 ···················· ···················· ····················· ···················· ···················· ····················
J 8527 hCoV-19/England/MILK-970406/2020|EPI_ISL_553119|20 ···················· ···················· ····················· ···················· ···················· ····················
J 8528 hCoV-19/England/MILK-970CC2/2020|EPI_ISL_553120|20 ···················· ···················· ····················· ···················· ···················· ····················
J 8529 hCoV-19/England/MILK-970828/2020|EPI_ISL_553121|20 ···················· ···················· ····················· ···················· ···················· ····················
J 8530 hCoV-19/England/MILK-970C86/2020|EPI_ISL_553123|20 ···················· ···················· ····················· ···················· ···················· ····················
J 8531 hCoV-19/England/MILK-970697/2020|EPI_ISL_553124|20 ···················· ···················· ····················· ···················· ···················· ····················
J 8532 hCoV-19/Wales/MILK-9712ED/2020|EPI_ISL_553125|2020 ···················· ···················· ····················· ···················· ···················· ····················
J 8533 hCoV-19/England/MILK-9706E2/2020|EPI_ISL_553126|20 ···················· ···················· ····················· ···················· ···················· ····················
J 8534 hCoV-19/Wales/MILK-97020C/2020|EPI_ISL_553127|2020 ···················· ···················· ····················· ···················· ···················· ····················
J 8535 hCoV-19/England/MILK-9704D9/2020|EPI_ISL_553130|20 ···················· ···················· ····················· ···················· ···················· ····················
J 8536 hCoV-19/Wales/MILK-971010/2020|EPI_ISL_553131|2020 ···················· ···················· ····················· ···················· ···················· ····················
J 8537 hCoV-19/England/MILK-9704AC/2020|EPI_ISL_553132|20 ···················· ···················· ····················· ···················· ···················· ····················
J 8538 hCoV-19/England/MILK-970F9C/2020|EPI_ISL_553133|20 ···················· ···················· ····················· ···················· ···················· ····················
J 8539 hCoV-19/England/MILK-9715B7/2020|EPI_ISL_553134|20 ···················· ···················· ····················· ···················· ···················· ····················
J 8540 hCoV-19/England/MILK-9709E9/2020|EPI_ISL_553135|20 ···················· ···················· ····················· ···················· ···················· ····················
J 8541 hCoV-19/England/MILK-970C4A/2020|EPI_ISL_553137|20 ···················· ···················· ····················· ···················· ···················· ····················
J 8542 hCoV-19/England/MILK-970D47/2020|EPI_ISL_553138|20 ···················· ···················· ····················· ···················· ···················· ····················
J 8543 hCoV-19/England/MILK-97055E/2020|EPI_ISL_553139|20 ···················· ···················· ····················· ···················· ···················· ····················
J 8544 hCoV-19/England/MILK-9717DF/2020|EPI_ISL_553140|20 ···················· ···················· ···········T········· ···················· ···················· ····················
J 8545 hCoV-19/Wales/MILK-9715E4/2020|EPI_ISL_553141|2020 ···················· ···················· ····················· ···················· ···················· ····················
J 8546 hCoV-19/England/MILK-9706B5/2020|EPI_ISL_553142|20 ···················· ···················· ····················· ···················· ···················· ····················
J 8547 hCoV-19/England/MILK-971687/2020|EPI_ISL_553143|20 ···················· ···················· ····················· ···················· ···················· ····················
J 8548 hCoV-19/England/MILK-970530/2020|EPI_ISL_553145|20 ···················· ···················· ····················· ···················· ···················· ····················
J 8549 hCoV-19/England/MILK-9705C7/2020|EPI_ISL_553146|20 ···················· ·A·················· ····················· ···················· ···················· ···A················
J 8550 hCoV-19/England/MILK-97064C/2020|EPI_ISL_553147|20 ···················· ···················· ····················· ···················· ···················· ····················
J 8551 hCoV-19/England/MILK-97103E/2020|EPI_ISL_553151|20 ···················· ···················· ····················· ···················· ···················· ····················
J 8552 hCoV-19/England/MILK-970512/2020|EPI_ISL_553152|20 ···················· ···················· ····················· ···················· ···················· ····················
J 8553 hCoV-19/England/MILK-970AD7/2020|EPI_ISL_553153|20 ···················· ···················· ····················· ···················· ···················· ····················
J 8554 hCoV-19/England/MILK-970864/2020|EPI_ISL_553154|20 ···················· ···················· ····················· ···················· ···················· ····················

| | | | | | | | | |
|---|---|---|---|---|---|---|---|---|
| J | 8555 | hCoV-19/England/MILK-970248/2020\|EPI_ISL_553155\|20 | ···················· | ·A·················· | ····················· | ···················· | ···················· | ···A················ |
| J | 8556 | hCoV-19/England/MILK-970970/2020\|EPI_ISL_553156\|20 | ···················· | ···················· | ····················· | ···················· | ···················· | ···················· |
| J | 8557 | hCoV-19/England/MILK-970EAE/2020\|EPI_ISL_553158\|20 | ···················· | ···················· | ····················· | ···················· | ···················· | ···················· |
| J | 8558 | hCoV-19/England/MILK-970794/2020\|EPI_ISL_553159\|20 | ···················· | ···················· | ····················· | ···················· | ···················· | ···················· |
| J | 8559 | hCoV-19/England/MILK-970D92/2020\|EPI_ISL_553161\|20 | ···················· | ·A·················· | ····················· | ···················· | ···················· | ···A················ |
| J | 8560 | hCoV-19/England/MILK-9705F4/2020\|EPI_ISL_553162\|20 | ···················· | ···················· | ····················· | ···················· | ···················· | ···················· |
| J | 8561 | hCoV-19/England/MILK-971089/2020\|EPI_ISL_553163\|20 | ···················· | ···················· | ····················· | ···················· | ···················· | ···················· |
| J | 8562 | hCoV-19/England/MILK-970EEA/2020\|EPI_ISL_553164\|20 | ···················· | ···················· | ····················· | ···················· | ···················· | ···················· |
| J | 8563 | hCoV-19/England/MILK-970381/2020\|EPI_ISL_553165\|20 | ···················· | ···················· | ····················· | ···················· | ···················· | ···················· |
| J | 8564 | hCoV-19/Wales/MILK-970B89/2020\|EPI_ISL_553166\|2020 | ···················· | ···················· | ····················· | ···················· | ···················· | ···················· |
| J | 8565 | hCoV-19/England/MILK-970BF2/2020\|EPI_ISL_553167\|20 | ···················· | ···················· | ····················· | ···················· | ···················· | ···················· |
| J | 8566 | hCoV-19/England/MILK-970D56/2020\|EPI_ISL_553168\|20 | ···················· | ···················· | ····················· | ···················· | ···················· | ···················· |
| J | 8567 | hCoV-19/England/MILK-97105C/2020\|EPI_ISL_553169\|20 | ···················· | ···················· | ···········T········· | ···················· | ···················· | ···················· |
| J | 8568 | hCoV-19/England/MILK-971168/2020\|EPI_ISL_553171\|20 | ···················· | ···················· | ····················· | ···················· | ···················· | ···················· |
| J | 8569 | hCoV-19/England/MILK-9713CC/2020\|EPI_ISL_553172\|20 | ···················· | ···················· | ····················· | ···················· | ···················· | ···················· |
| J | 8570 | hCoV-19/England/MILK-9708CE/2020\|EPI_ISL_553173\|20 | ···················· | ···················· | ····················· | ···················· | ···················· | ···················· |
| J | 8571 | hCoV-19/England/MILK-970336/2020\|EPI_ISL_553174\|20 | ···················· | ···················· | ····················· | ···················· | ···················· | ···················· |
| J | 8572 | hCoV-19/England/MILK-971292/2020\|EPI_ISL_553176\|20 | ···················· | ···················· | ····················· | ···················· | ···················· | ···················· |
| J | 8573 | hCoV-19/England/MILK-9715A8/2020\|EPI_ISL_553177\|20 | ···················· | ···················· | ····················· | ···················· | ···················· | ···················· |
| J | 8574 | hCoV-19/England/MILK-970DFC/2020\|EPI_ISL_553178\|20 | ···················· | ···················· | ····················· | ···················· | ···················· | ···················· |
| J | 8575 | hCoV-19/England/MILK-970AE6/2020\|EPI_ISL_553179\|20 | ···················· | ···················· | ····················· | ···················· | ···················· | ···················· |
| J | 8576 | hCoV-19/England/MILK-9705A9/2020\|EPI_ISL_553180\|20 | ···················· | ·A·················· | ····················· | ···················· | ···················· | ···A················ |
| J | 8577 | hCoV-19/England/MILK-9707A3/2020\|EPI_ISL_553181\|20 | ···················· | ·A·················· | ····················· | ···················· | ···················· | ···A················ |
| J | 8578 | hCoV-19/England/MILK-970BB6/2020\|EPI_ISL_553184\|20 | ···················· | ···················· | ····················· | ···················· | ···················· | ···················· |
| J | 8579 | hCoV-19/England/MILK-97070D/2020\|EPI_ISL_553185\|20 | ···················· | ···················· | ····················· | ···················· | ···················· | ···················· |
| J | 8580 | hCoV-19/England/MILK-970BA7/2020\|EPI_ISL_553188\|20 | ···················· | ···················· | ····················· | ···················· | ······T············· | ···················· |
| J | 8581 | hCoV-19/England/MILK-971845/2020\|EPI_ISL_553189\|20 | ···················· | ·A·················· | ····················· | ···················· | ···················· | ···················· |
| J | 8582 | hCoV-19/England/MILK-971308/2020\|EPI_ISL_553190\|20 | ···················· | ···················· | ····················· | ···················· | ···················· | ···················· |
| J | 8583 | hCoV-19/England/MILK-9710B6/2020\|EPI_ISL_553191\|20 | ···················· | ·A·················· | ····················· | ···················· | ···················· | ···A················ |
| J | 8584 | hCoV-19/England/MILK-9704F7/2020\|EPI_ISL_553192\|20 | ···················· | ···················· | ····················· | ···················· | ······T············· | ···················· |
| J | 8585 | hCoV-19/England/MILK-9706C4/2020\|EPI_ISL_553194\|20 | ···················· | ···················· | ····················· | ···················· | ···················· | ···················· |
| J | 8586 | hCoV-19/Wales/MILK-971001/2020\|EPI_ISL_553195\|2020 | ···················· | ···················· | ····················· | ···················· | ···················· | ···················· |
| J | 8587 | hCoV-19/England/MILK-971283/2020\|EPI_ISL_553196\|20 | ···················· | ···················· | ····················· | ···················· | ···················· | ···················· |
| J | 8588 | hCoV-19/England/MILK-971256/2020\|EPI_ISL_553197\|20 | ···················· | ···················· | ····················· | ···················· | ···················· | ···················· |
| J | 8589 | hCoV-19/England/MILK-970E08/2020\|EPI_ISL_553198\|20 | ···················· | ···················· | ····················· | ···················· | ···················· | ···················· |
| J | 8590 | hCoV-19/England/ALDP-955892/2020\|EPI_ISL_554446\|20 | ···················· | ···················· | ····················· | ···················· | ···················· | ···················· |
| J | 8591 | hCoV-19/England/ALDP-952FAF/2020\|EPI_ISL_554447\|20 | ···················· | ···················· | ····················· | ···················· | ···················· | ···················· |
| J | 8592 | hCoV-19/England/ALDP-95092B/2020\|EPI_ISL_554448\|20 | ···················· | ···················· | ····················· | ···················· | ···················· | ···················· |
| J | 8593 | hCoV-19/England/ALDP-95166F/2020\|EPI_ISL_554449\|20 | ···················· | ···················· | ····················· | ···················· | ···················· | ···················· |
| J | 8594 | hCoV-19/England/ALDP-953C98/2020\|EPI_ISL_554450\|20 | ···················· | ···················· | ····················· | ···················· | ···················· | ···················· |
| J | 8595 | hCoV-19/England/ALDP-9511B9/2020\|EPI_ISL_554451\|20 | ···················· | ···················· | ····················· | ···················· | ···················· | ···················· |
| J | 8596 | hCoV-19/England/ALDP-9532B4/2020\|EPI_ISL_554452\|20 | ···················· | ···················· | ····················· | ···················· | ···················· | ···················· |
| J | 8597 | hCoV-19/England/ALDP-955847/2020\|EPI_ISL_554453\|20 | ···················· | ···················· | ····················· | ···················· | ···················· | ···················· |
| J | 8598 | hCoV-19/England/ALDP-953FDB/2020\|EPI_ISL_554454\|20 | ···················· | ···················· | ····················· | ···················· | ···················· | ···················· |
| J | 8599 | hCoV-19/England/ALDP-950A82/2020\|EPI_ISL_554455\|20 | ···················· | ···················· | ····················· | ···················· | ···················· | ···················· |
| J | 8600 | hCoV-19/England/ALDP-9558FC/2020\|EPI_ISL_554456\|20 | ···················· | ···················· | ····················· | ···················· | ···················· | ···················· |
| J | 8601 | hCoV-19/England/ALDP-953490/2020\|EPI_ISL_554457\|20 | ···················· | ···················· | ····················· | ···················· | ···················· | ···················· |
| J | 8602 | hCoV-19/England/ALDP-9534AF/2020\|EPI_ISL_554458\|20 | ···················· | ···················· | ····················· | ···················· | ···················· | ···················· |
| J | 8603 | hCoV-19/England/ALDP-950D10/2020\|EPI_ISL_554460\|20 | ···················· | ···················· | ····················· | ···················· | ···················· | ···················· |
| J | 8604 | hCoV-19/England/ALDP-951650/2020\|EPI_ISL_554461\|20 | ···················· | ···················· | ····················· | ···················· | ···················· | ···················· |
| J | 8605 | hCoV-19/Wales/ALDP-95320F/2020\|EPI_ISL_554462\|2020 | ···················· | ···················· | ····················· | ···················· | ···················· | ···················· |
| J | 8606 | hCoV-19/England/ALDP-950BDA/2020\|EPI_ISL_554463\|20 | ···················· | ···················· | ····················· | ···················· | ···················· | ···················· |
| J | 8607 | hCoV-19/England/ALDP-950E0E/2020\|EPI_ISL_554464\|20 | ···················· | ·A·················· | ····················· | ···················· | ···················· | ···················· |
| J | 8608 | hCoV-19/England/ALDP-9531A8/2020\|EPI_ISL_554465\|20 | ···················· | ···················· | ····················· | ···················· | ···················· | ···················· |
| J | 8609 | hCoV-19/England/ALDP-951614/2020\|EPI_ISL_554466\|20 | ···················· | ···················· | ····················· | ···················· | ···················· | ···················· |
| J | 8610 | hCoV-19/England/ALDP-95124D/2020\|EPI_ISL_554467\|20 | ···················· | ···················· | ····················· | ···················· | ···················· | ···················· |
| J | 8611 | hCoV-19/England/ALDP-950ADD/2020\|EPI_ISL_554468\|20 | ···················· | ···················· | ····················· | ···················· | ···················· | ···················· |
| J | 8612 | hCoV-19/England/ALDP-95086A/2020\|EPI_ISL_554469\|20 | ···················· | ···················· | ····················· | ···················· | ···················· | ···················· |
| J | 8613 | hCoV-19/England/ALDP-95548F/2020\|EPI_ISL_554471\|20 | ···················· | ·A·················· | ····················· | ···················· | ···················· | ···················· |
| J | 8614 | hCoV-19/England/ALDP-955829/2020\|EPI_ISL_554472\|20 | ···················· | ···················· | ····················· | ···················· | ···················· | ···················· |
| J | 8615 | hCoV-19/England/ALDP-953120/2020\|EPI_ISL_554473\|20 | ···················· | ···················· | ····················· | ···················· | ···················· | ···················· |
| J | 8616 | hCoV-19/England/ALDP-953788/2020\|EPI_ISL_554474\|20 | ···················· | ···················· | ····················· | ···················· | ···················· | ···················· |
| J | 8617 | hCoV-19/England/ALDP-950DB6/2020\|EPI_ISL_554476\|20 | ···················· | ···················· | ····················· | ···················· | ···················· | ···················· |
| J | 8618 | hCoV-19/England/ALDP-951438/2020\|EPI_ISL_554477\|20 | ···················· | ·A·················· | ····················· | ···················· | ···················· | ···················· |
| J | 8619 | hCoV-19/England/ALDP-9514DE/2020\|EPI_ISL_554480\|20 | ···················· | ···················· | ····················· | ···················· | ···················· | ···················· |
| J | 8620 | hCoV-19/England/ALDP-95169C/2020\|EPI_ISL_554481\|20 | ···················· | ···················· | ····················· | ···················· | ···················· | ···················· |
| J | 8621 | hCoV-19/England/ALDP-953F80/2020\|EPI_ISL_554482\|20 | ···················· | ···················· | ····················· | ···················· | ···················· | ···················· |
| J | 8622 | hCoV-19/England/ALDP-953ACB/2020\|EPI_ISL_554483\|20 | ···················· | ···················· | ····················· | ···················· | ···················· | ···················· |
| J | 8623 | hCoV-19/England/ALDP-955759/2020\|EPI_ISL_554485\|20 | ···················· | ···················· | ····················· | ···················· | ···················· | ···················· |
| J | 8624 | hCoV-19/England/ALDP-953E65/2020\|EPI_ISL_554486\|20 | ···················· | ···················· | ····················· | ···················· | ···················· | ···················· |
| J | 8625 | hCoV-19/England/ALDP-950DC5/2020\|EPI_ISL_554488\|20 | ···················· | ···················· | ····················· | ···················· | ···················· | ···················· |
| J | 8626 | hCoV-19/England/ALDP-950A19/2020\|EPI_ISL_554489\|20 | ···················· | ···················· | ····················· | ···················· | ···················· | ···················· |
| J | 8627 | hCoV-19/England/ALDP-953384/2020\|EPI_ISL_554490\|20 | ···················· | ···················· | ····················· | ···················· | ···················· | ···················· |
| J | 8628 | hCoV-19/England/ALDP-95380D/2020\|EPI_ISL_554493\|20 | ···················· | ···················· | ····················· | ···················· | ···················· | ···················· |
| J | 8629 | hCoV-19/England/ALDP-95365E/2020\|EPI_ISL_554494\|20 | ···················· | ···················· | ····················· | ···················· | ···················· | ···················· |
| J | 8630 | hCoV-19/England/ALDP-95313F/2020\|EPI_ISL_554495\|20 | ···················· | ···················· | ····················· | ···················· | ···················· | ···················· |
| J | 8631 | hCoV-19/England/ALDP-95369A/2020\|EPI_ISL_554496\|20 | ···················· | ···················· | ····················· | ···················· | ···················· | ···················· |
| J | 8632 | hCoV-19/England/ALDP-9508D3/2020\|EPI_ISL_554499\|20 | ···················· | ···················· | ····················· | ···················· | ···················· | ···················· |
| J | 8633 | hCoV-19/England/ALDP-94FAC2/2020\|EPI_ISL_555777\|20 | ···················· | ···················· | ····················· | ···················· | ···················· | ···················· |
| J | 8634 | hCoV-19/England/ALDP-94F7BC/2020\|EPI_ISL_555779\|20 | ···················· | ···················· | ····················· | ···················· | ···················· | ···················· |

| | | | | | | | | |
|---|---|---|---|---|---|---|---|---|
| J | 8635 | hCoV-19/England/ALDP-94B871/2020\|EPI_ISL_555780\|20 | ···················· | ···················· | ···················· | ···················· | ···················· | ···················· |
| J | 8636 | hCoV-19/England/ALDP-94A9BB/2020\|EPI_ISL_555782\|20 | ···················· | ···················· | ···················· | ···················· | ···················· | ···················· |
| J | 8637 | hCoV-19/England/ALDP-94BBB4/2020\|EPI_ISL_555784\|20 | ···················· | ···················· | ··········T········· | ···················· | ···················· | ···················· |
| J | 8638 | hCoV-19/England/ALDP-94B774/2020\|EPI_ISL_555785\|20 | ···················· | ···················· | ···················· | ···················· | ···················· | ···················· |
| J | 8639 | hCoV-19/England/ALDP-94F804/2020\|EPI_ISL_555786\|20 | ···················· | ···················· | ···················· | ···················· | ···················· | ···················· |
| J | 8640 | hCoV-19/England/ALDP-94B12B/2020\|EPI_ISL_555787\|20 | ···················· | ···················· | ···················· | ···················· | ···················· | ···················· |
| J | 8641 | hCoV-19/England/ALDP-94B96F/2020\|EPI_ISL_555788\|20 | ···················· | ···················· | ···················· | ···················· | ···················· | ···················· |
| J | 8642 | hCoV-19/England/ALDP-94F813/2020\|EPI_ISL_555789\|20 | ···················· | ···················· | ···················· | ···················· | ···················· | ···················· |
| J | 8643 | hCoV-19/England/ALDP-94B932/2020\|EPI_ISL_555790\|20 | ···················· | ···················· | ···················· | ···················· | ···················· | ···················· |
| J | 8644 | hCoV-19/England/ALDP-94FEC6/2020\|EPI_ISL_555791\|20 | ···················· | ···················· | ···················· | ···················· | ···················· | ···················· |
| J | 8645 | hCoV-19/England/ALDP-94BB96/2020\|EPI_ISL_555792\|20 | ···················· | ···················· | ···················· | ···················· | ···················· | ···················· |
| J | 8646 | hCoV-19/England/ALDP-94BA4E/2020\|EPI_ISL_555793\|20 | ···················· | ···················· | ···················· | ···················· | ···················· | ···················· |
| J | 8647 | hCoV-19/England/ALDP-94A63C/2020\|EPI_ISL_555795\|20 | ···················· | ···················· | ···················· | ···················· | ···················· | ···················· |
| J | 8648 | hCoV-19/England/ALDP-94BB78/2020\|EPI_ISL_555796\|20 | ···················· | ···················· | ···················· | ···················· | ···················· | ···················· |
| J | 8649 | hCoV-19/England/ALDP-94FAD1/2020\|EPI_ISL_555797\|20 | ···················· | ···················· | ···················· | ···················· | ···················· | ···················· |
| J | 8650 | hCoV-19/Wales/ALDP-94F725/2020\|EPI_ISL_555798\|2020 | ···················· | ···················· | ···················· | ···················· | ···················· | ···················· |
| J | 8651 | hCoV-19/England/ALDP-94A65A/2020\|EPI_ISL_555799\|20 | ···················· | ···················· | ···················· | ···················· | ···················· | ···················· |
| J | 8652 | hCoV-19/England/ALDP-68CE1B/2020\|EPI_ISL_558002\|20 | ···················· | ···················· | ···················· | ···················· | ···················· | ···················· |
| J | 8653 | hCoV-19/England/ALDP-66DFA4/2020\|EPI_ISL_558003\|20 | ···················· | ···················· | ···················· | ···················· | ···················· | ···················· |
| J | 8654 | hCoV-19/England/ALDP-6473FF/2020\|EPI_ISL_558004\|20 | ···················· | ···················· | ···················· | ···················· | ···················· | ···················· |
| J | 8655 | hCoV-19/England/ALDP-65781B/2020\|EPI_ISL_558005\|20 | ···················· | ···················· | ···················· | ···················· | ···················· | ···················· |
| J | 8656 | hCoV-19/England/ALDP-66E0A0/2020\|EPI_ISL_558007\|20 | ···················· | ···················· | ···················· | ···················· | ···················· | ···················· |
| J | 8657 | hCoV-19/England/ALDP-645203/2020\|EPI_ISL_558008\|20 | ···················· | ···················· | ···················· | ···················· | ···················· | ···················· |
| J | 8658 | hCoV-19/England/ALDP-68B90C/2020\|EPI_ISL_558009\|20 | ···················· | ···················· | ···················· | ···················· | ···················· | ···················· |
| J | 8659 | hCoV-19/England/MILK-4A19A8/2020\|EPI_ISL_559300\|20 | ···················· | ···················· | ···················· | ···················· | ···················· | ···················· |
| J | 8660 | hCoV-19/England/ALDP-4AD21A/2020\|EPI_ISL_559301\|20 | ···················· | ···················· | ···················· | ···················· | ···················· | ···················· |
| J | 8661 | hCoV-19/England/ALDP-49AD1F/2020\|EPI_ISL_559302\|20 | ···················· | ···················· | ···················· | ···················· | ···················· | ···················· |
| J | 8662 | hCoV-19/England/ALDP-4AD283/2020\|EPI_ISL_559303\|20 | ···················· | ···················· | ···················· | ···················· | ···················· | ···················· |
| J | 8663 | hCoV-19/England/ALDP-496B19/2020\|EPI_ISL_559304\|20 | ···················· | ···················· | ···················· | ···················· | ···················· | ···················· |
| J | 8664 | hCoV-19/England/ALDP-49B464/2020\|EPI_ISL_559305\|20 | ···················· | ···················· | ···················· | ···················· | ···················· | ···················· |
| J | 8665 | hCoV-19/England/ALDP-4AF3D9/2020\|EPI_ISL_559306\|20 | ···················· | ···················· | ···················· | ···················· | ···················· | ···················· |
| J | 8666 | hCoV-19/England/ALDP-4AECB1/2020\|EPI_ISL_559307\|20 | ···················· | ·A·················· | ···················· | ···················· | ···················· | ···················· |
| J | 8667 | hCoV-19/England/MILK-4A0A79/2020\|EPI_ISL_559319\|20 | ···················· | ···················· | ···················· | ···················· | ···················· | ···················· |
| J | 8668 | hCoV-19/England/MILK-4A1665/2020\|EPI_ISL_559320\|20 | ···················· | ···················· | ···················· | ···················· | ···················· | ···················· |
| J | 8669 | hCoV-19/England/ALDP-49B0F7/2020\|EPI_ISL_559321\|20 | ···················· | ···················· | ···················· | ···················· | ···················· | ···················· |
| J | 8670 | hCoV-19/England/ALDP-49BB50/2020\|EPI_ISL_559322\|20 | ···················· | ···················· | ···················· | ···················· | ···················· | ···················· |
| J | 8671 | hCoV-19/England/ALDP-49B19A/2020\|EPI_ISL_559323\|20 | ···················· | ···················· | ···················· | ···················· | ···················· | ···················· |
| J | 8672 | hCoV-19/England/ALDP-4AD775/2020\|EPI_ISL_559324\|20 | ···················· | ···················· | ···················· | ···················· | ···················· | ···················· |
| J | 8673 | hCoV-19/England/ALDP-4976EA/2020\|EPI_ISL_559325\|20 | ···················· | ···················· | ···················· | ···················· | ···················· | ···················· |
| J | 8674 | hCoV-19/England/MILK-49FE44/2020\|EPI_ISL_559326\|20 | ···················· | ···················· | ···················· | ···················· | ···················· | ···················· |
| J | 8675 | hCoV-19/England/ALDP-49BA80/2020\|EPI_ISL_559327\|20 | ···················· | ···················· | ···················· | ···················· | ···················· | ···················· |
| J | 8676 | hCoV-19/England/ALDP-49B5E9/2020\|EPI_ISL_559328\|20 | ···················· | ···················· | ···················· | ···················· | ···················· | ···················· |
| J | 8677 | hCoV-19/England/ALDP-49BE84/2020\|EPI_ISL_559329\|20 | ···················· | ···················· | ···················· | ···················· | ···················· | ···················· |
| J | 8678 | hCoV-19/England/ALDP-49B525/2020\|EPI_ISL_559330\|20 | ···················· | ···················· | ···················· | ···················· | ···················· | ···················· |
| J | 8679 | hCoV-19/England/MILK-496593/2020\|EPI_ISL_559331\|20 | ···················· | ···················· | ···················· | ···················· | ···················· | ···················· |
| J | 8680 | hCoV-19/England/ALDP-49BDD2/2020\|EPI_ISL_559332\|20 | ···················· | ···················· | ···················· | ···················· | ···················· | ···················· |
| J | 8681 | hCoV-19/England/ALDP-4AF166/2020\|EPI_ISL_559333\|20 | ···················· | ···················· | ···················· | ···················· | ···················· | ···················· |
| J | 8682 | hCoV-19/England/ALDP-4966BE/2020\|EPI_ISL_559334\|20 | ···················· | ···················· | ···················· | ···················· | ···················· | ···················· |
| J | 8683 | hCoV-19/England/MILK-49FFC9/2020\|EPI_ISL_559335\|20 | ···················· | ···················· | ···················· | ···················· | ···················· | ···················· |
| J | 8684 | hCoV-19/England/ALDP-496E6B/2020\|EPI_ISL_559336\|20 | ···················· | ···················· | ···················· | ···················· | ···················· | ···················· |
| J | 8685 | hCoV-19/England/ALDP-496B37/2020\|EPI_ISL_559338\|20 | ···················· | ···················· | ···················· | ···················· | ···················· | ···················· |
| J | 8686 | hCoV-19/England/ALDP-4AD2DE/2020\|EPI_ISL_559339\|20 | ···················· | ···················· | ···················· | ···················· | ···················· | ···················· |
| J | 8687 | hCoV-19/Wales/PHWC-16B512/2020\|EPI_ISL_560300\|2020 | ···················· | ···················· | ···················· | ···················· | ···················· | ···················· |
| J | 8688 | hCoV-19/Wales/PHWC-16B54F/2020\|EPI_ISL_560301\|2020 | ···················· | ···················· | ···················· | ···················· | ···················· | ···················· |
| J | 8689 | hCoV-19/Wales/PHWC-16B55E/2020\|EPI_ISL_560302\|2020 | ···················· | ···················· | ···················· | ···················· | ···················· | ···················· |
| J | 8690 | hCoV-19/Wales/PHWC-16B57C/2020\|EPI_ISL_560304\|2020 | ···················· | ···················· | ···················· | ···················· | ···················· | ···················· |
| J | 8691 | hCoV-19/Wales/QEUH-99630D/2020\|EPI_ISL_560305\|2020 | ···················· | ···················· | ···················· | ···················· | ···················· | ···················· |
| J | 8692 | hCoV-19/Wales/QEUH-99C264/2020\|EPI_ISL_560306\|2020 | ···················· | ···················· | ···················· | ···················· | ···················· | ···················· |
| J | 8693 | hCoV-19/Russia/SVE-RII-MH182S/2020\|EPI_ISL_560308\| | ···················· | ···················· | ···················· | ···················· | ···················· | ···················· |
| J | 8694 | hCoV-19/Russia/SVE-RII-MH184S/2020\|EPI_ISL_560309\| | ···················· | ···················· | ···················· | ···················· | ···················· | ···················· |
| J | 8695 | hCoV-19/Russia/SVE-RII-MH187S/2020\|EPI_ISL_560310\| | ···················· | ···················· | ···················· | ···················· | ···················· | ···················· |
| J | 8696 | hCoV-19/Russia/SVE-RII-MH197S/2020\|EPI_ISL_560311\| | ···················· | ···················· | ···················· | ···················· | ···················· | ···················· |
| J | 8697 | hCoV-19/Russia/SVE-RII-MH198S/2020\|EPI_ISL_560312\| | ···················· | ···················· | ···················· | ···················· | ···················· | ···················· |
| J | 8698 | hCoV-19/Russia/SVE-RII-MH199S/2020\|EPI_ISL_560313\| | ···················· | ···················· | ···················· | ···················· | ···················· | ···················· |
| J | 8699 | hCoV-19/Russia/SVE-RII-MH200S/2020\|EPI_ISL_560314\| | ···················· | ···················· | ···················· | ···················· | ···················· | ···················· |
| J | 8700 | hCoV-19/Russia/SVE-RII-MH202S/2020\|EPI_ISL_560315\| | ···················· | ···················· | ···················· | ···················· | ···················· | ···················· |
| J | 8701 | hCoV-19/Russia/SVE-RII-MH210S/2020\|EPI_ISL_560316\| | ···················· | ···················· | ···················· | ···················· | ···················· | ···················· |
| J | 8702 | hCoV-19/Russia/SVE-RII-MH231S/2020\|EPI_ISL_560317\| | ···················· | ···················· | ···················· | ···················· | ···················· | ···················· |
| J | 8703 | hCoV-19/India/HR-IMT-AE21/2020\|EPI_ISL_560318\|2020 | ···················· | ···················· | ···················· | ···················· | ···················· | ···················· |
| J | 8704 | hCoV-19/India/UT-IMT-AN32/2020\|EPI_ISL_560319\|2020 | ···················· | ·A·················· | ···················· | ···················· | ···················· | ···A················ |
| J | 8705 | hCoV-19/England/MILK-99D0B4/2020\|EPI_ISL_549926\|20 | ···················· | ···················· | ···················· | ···················· | ···················· | ···················· |
| J | 8706 | hCoV-19/England/MILK-99CDFA/2020\|EPI_ISL_549929\|20 | ···················· | ···················· | ···················· | ···················· | ···················· | ···················· |
| J | 8707 | hCoV-19/England/MILK-9AA03C/2020\|EPI_ISL_550910\|20 | ···················· | ···················· | ···················· | ···················· | ···················· | ···················· |
| J | 8708 | hCoV-19/England/MILK-9AA333/2020\|EPI_ISL_550911\|20 | ···················· | ·A·················· | ···················· | ···················· | ···················· | ···A················ |
| J | 8709 | hCoV-19/England/MILK-9AAD08/2020\|EPI_ISL_550912\|20 | ···················· | ···················· | ···················· | ···················· | ···················· | ···················· |
| J | 8710 | hCoV-19/England/MILK-9AA773/2020\|EPI_ISL_550913\|20 | ···················· | ···················· | ···················· | ···················· | ···················· | ···················· |
| J | 8711 | hCoV-19/Wales/MILK-9AA61C/2020\|EPI_ISL_550914\|2020 | ···················· | ···················· | ···················· | ···················· | ···················· | ···················· |
| J | 8712 | hCoV-19/England/MILK-9AACCF/2020\|EPI_ISL_550915\|20 | ···················· | ···················· | ···················· | ···················· | ···················· | ···················· |
| J | 8713 | hCoV-19/England/MILK-9AA834/2020\|EPI_ISL_550918\|20 | ···················· | ···················· | ···················· | ···················· | ···················· | ···················· |
| J | 8714 | hCoV-19/England/MILK-9AA8BC/2020\|EPI_ISL_550919\|20 | ···················· | C··················· | ···················· | ···················· | ···················· | ···················· |

| | | | | | | | |
|---|---|---|---|---|---|---|---|
| J | 8715 hCoV-19/England/MILK-9709DA/2020\|EPI_ISL_553200\|20 | ···················· | ···················· | ···················· | ···················· | ···················· | ···················· |
| J | 8716 hCoV-19/England/MILK-970CE0/2020\|EPI_ISL_553201\|20 | ···················· | ·A·················· | ···················· | ···················· | ···················· | ···A················ |
| J | 8717 hCoV-19/England/MILK-9706A6/2020\|EPI_ISL_553203\|20 | ···················· | ···················· | ···················· | ···················· | ···················· | ···················· |
| J | 8718 hCoV-19/England/MILK-97112C/2020\|EPI_ISL_553204\|20 | ···················· | ···················· | ···················· | ···················· | ···················· | ···················· |
| J | 8719 hCoV-19/England/MILK-970F41/2020\|EPI_ISL_553205\|20 | ···················· | ···················· | ···················· | ···················· | ···················· | ···················· |
| J | 8720 hCoV-19/England/MILK-97022A/2020\|EPI_ISL_553207\|20 | ···················· | ···················· | ···················· | ···················· | ···················· | ···················· |
| J | 8721 hCoV-19/England/MILK-970AF5/2020\|EPI_ISL_553208\|20 | ···················· | ·A·················· | ···················· | ···················· | ···················· | ···A················ |
| J | 8722 hCoV-19/England/MILK-97153F/2020\|EPI_ISL_553209\|20 | ···················· | ···················· | ···················· | ···················· | ···················· | ···················· |
| J | 8723 hCoV-19/England/MILK-9707D0/2020\|EPI_ISL_553210\|20 | ···················· | ···················· | ···················· | ···················· | ···················· | ···················· |
| J | 8724 hCoV-19/England/MILK-970758/2020\|EPI_ISL_553211\|20 | ···················· | ···················· | ···················· | ···················· | ···················· | ···················· |
| J | 8725 hCoV-19/Wales/MILK-970E26/2020\|EPI_ISL_553212\|2020 | ···················· | ···················· | ···················· | ···················· | ···················· | ···················· |
| J | 8726 hCoV-19/England/MILK-97047F/2020\|EPI_ISL_553213\|20 | ···················· | ···················· | ···················· | ···················· | ···················· | ···················· |
| J | 8727 hCoV-19/England/MILK-971818/2020\|EPI_ISL_553214\|20 | ···················· | ···················· | ···················· | ···················· | ···················· | ···················· |
| J | 8728 hCoV-19/England/MILK-97114A/2020\|EPI_ISL_553216\|20 | ···················· | ···················· | ···················· | ···················· | ···················· | ···················· |
| J | 8729 hCoV-19/England/MILK-971371/2020\|EPI_ISL_553217\|20 | ···················· | ···················· | ···················· | ···················· | ···················· | ···················· |
| J | 8730 hCoV-19/England/MILK-970318/2020\|EPI_ISL_553218\|20 | ···················· | ·A·················· | ···················· | ···················· | ···················· | ···················· |
| J | 8731 hCoV-19/Wales/MILK-970B6B/2020\|EPI_ISL_553219\|2020 | ···················· | ···················· | ···················· | ···················· | ···················· | ···················· |
| J | 8732 hCoV-19/England/MILK-971344/2020\|EPI_ISL_553220\|20 | ···················· | ···················· | ···················· | ···················· | ······T············· | ···················· |
| J | 8733 hCoV-19/Wales/MILK-970B7A/2020\|EPI_ISL_553221\|2020 | ···················· | ···················· | ···················· | ···················· | ···················· | ···················· |
| J | 8734 hCoV-19/England/MILK-97066A/2020\|EPI_ISL_553222\|20 | ···················· | ···················· | ···················· | ···················· | ···················· | ···················· |
| J | 8735 hCoV-19/Wales/MILK-9712DE/2020\|EPI_ISL_553223\|2020 | ···················· | ···················· | ···················· | ···················· | ···················· | ···················· |
| J | 8736 hCoV-19/England/MILK-971247/2020\|EPI_ISL_553224\|20 | ···················· | ···················· | ···················· | ···················· | ···················· | ···················· |
| J | 8737 hCoV-19/England/MILK-970BE3/2020\|EPI_ISL_553225\|20 | ···················· | ···················· | ···················· | ···················· | ···················· | ···················· |
| J | 8738 hCoV-19/England/MILK-970A04/2020\|EPI_ISL_553226\|20 | ···················· | ···················· | ···················· | ···················· | ···················· | ···············A···· |
| J | 8739 hCoV-19/England/MILK-970266/2020\|EPI_ISL_553227\|20 | ···················· | ···················· | ···················· | ···················· | ···················· | ···················· |
| J | 8740 hCoV-19/England/MILK-9711FF/2020\|EPI_ISL_553228\|20 | ···················· | ···················· | ···················· | ···················· | ···················· | ···················· |
| J | 8741 hCoV-19/England/MILK-9708FB/2020\|EPI_ISL_553229\|20 | ···················· | ···················· | ···················· | ···················· | ···················· | ···················· |
| J | 8742 hCoV-19/England/MILK-970415/2020\|EPI_ISL_553230\|20 | ···················· | ···················· | ···················· | ···················· | ···················· | ···················· |
| J | 8743 hCoV-19/England/MILK-9716C3/2020\|EPI_ISL_553232\|20 | ···················· | ···················· | ···················· | ···················· | ···················· | ···················· |
| J | 8744 hCoV-19/England/MILK-971739/2020\|EPI_ISL_553233\|20 | ···················· | ···················· | ···················· | ···················· | ···················· | ···················· |
| J | 8745 hCoV-19/England/ALDP-9511F5/2020\|EPI_ISL_554500\|20 | ···················· | ···················· | ···················· | ···················· | ···················· | ···················· |
| J | 8746 hCoV-19/England/ALDP-952FDC/2020\|EPI_ISL_554501\|20 | ···················· | ···················· | ···················· | ···················· | ···················· | ···················· |
| J | 8747 hCoV-19/England/ALDP-9533A2/2020\|EPI_ISL_554502\|20 | ···················· | ···················· | ···················· | ···················· | ···················· | ···················· |
| J | 8748 hCoV-19/England/ALDP-9534DC/2020\|EPI_ISL_554503\|20 | ···················· | ···················· | ···················· | ···················· | ···················· | ···················· |
| J | 8749 hCoV-19/England/ALDP-9507E5/2020\|EPI_ISL_554504\|20 | ···················· | ···················· | ···················· | ···················· | ···················· | ···················· |
| J | 8750 hCoV-19/England/ALDP-95081F/2020\|EPI_ISL_554505\|20 | ···················· | ···················· | ···················· | ···················· | ···················· | ···················· |
| J | 8751 hCoV-19/England/ALDP-951298/2020\|EPI_ISL_554507\|20 | ···················· | ···················· | ···················· | ···················· | ···················· | ···················· |
| J | 8752 hCoV-19/England/ALDP-954277/2020\|EPI_ISL_554508\|20 | ···················· | ···················· | ···················· | ···················· | ···················· | ···················· |
| J | 8753 hCoV-19/England/ALDP-951544/2020\|EPI_ISL_554509\|20 | ···················· | ···················· | ···················· | ···················· | ···················· | ···················· |
| J | 8754 hCoV-19/England/ALDP-9534CD/2020\|EPI_ISL_554511\|20 | ···················· | ···················· | ···················· | ···················· | ···················· | ···················· |
| J | 8755 hCoV-19/England/ALDP-950DD4/2020\|EPI_ISL_554512\|20 | ···················· | ···················· | ···················· | ···················· | ···················· | ···················· |
| J | 8756 hCoV-19/England/ALDP-952F63/2020\|EPI_ISL_554513\|20 | ···················· | ···················· | ···················· | ···················· | ···················· | ···················· |
| J | 8757 hCoV-19/England/ALDP-950C31/2020\|EPI_ISL_554514\|20 | ···················· | ···················· | ···················· | ···················· | ···················· | ···················· |
| J | 8758 hCoV-19/England/ALDP-950B25/2020\|EPI_ISL_554515\|20 | ···················· | ·A·················· | ···················· | ···················· | ···················· | ···················· |
| J | 8759 hCoV-19/England/ALDP-950D01/2020\|EPI_ISL_554516\|20 | ···················· | ···················· | ···················· | ···················· | ···················· | ···················· |
| J | 8760 hCoV-19/England/ALDP-955935/2020\|EPI_ISL_554517\|20 | ···················· | ···················· | ···················· | ···················· | ···················· | ···················· |
| J | 8761 hCoV-19/England/ALDP-950CD7/2020\|EPI_ISL_554518\|20 | ···················· | ···················· | ···················· | ···················· | ···················· | ···················· |
| J | 8762 hCoV-19/England/ALDP-953E83/2020\|EPI_ISL_554520\|20 | ···················· | ···················· | ···················· | ···················· | ···················· | ···················· |
| J | 8763 hCoV-19/England/ALDP-950C40/2020\|EPI_ISL_554521\|20 | ···················· | ···················· | ···················· | ···················· | ···················· | ···················· |
| J | 8764 hCoV-19/England/ALDP-9531B7/2020\|EPI_ISL_554522\|20 | ···················· | ···················· | ···················· | ···················· | ···················· | ···················· |
| J | 8765 hCoV-19/England/ALDP-95581A/2020\|EPI_ISL_554523\|20 | ···················· | ···················· | ···················· | ···················· | ···················· | ···················· |
| J | 8766 hCoV-19/England/ALDP-95123E/2020\|EPI_ISL_554524\|20 | ···················· | ···················· | ···················· | ···················· | ···················· | ···················· |
| J | 8767 hCoV-19/England/ALDP-953CA7/2020\|EPI_ISL_554525\|20 | ···················· | ···················· | ···················· | ···················· | ···················· | ···················· |
| J | 8768 hCoV-19/England/ALDP-950BBC/2020\|EPI_ISL_554527\|20 | ···················· | ···················· | ···················· | ···················· | ···················· | ···················· |
| J | 8769 hCoV-19/England/ALDP-950B9E/2020\|EPI_ISL_554529\|20 | ···················· | ···················· | ···················· | ···················· | ···················· | ···················· |
| J | 8770 hCoV-19/England/ALDP-95564D/2020\|EPI_ISL_554530\|20 | ···················· | ···················· | ···················· | ···················· | ···················· | ···················· |
| J | 8771 hCoV-19/England/ALDP-953ABC/2020\|EPI_ISL_554531\|20 | ···················· | ···················· | ···················· | ···················· | ···················· | ···················· |
| J | 8772 hCoV-19/England/ALDP-953E74/2020\|EPI_ISL_554532\|20 | ···················· | ···················· | ···················· | ···················· | ···················· | ···················· |
| J | 8773 hCoV-19/England/ALDP-955443/2020\|EPI_ISL_554533\|20 | ···················· | ···················· | ···················· | ···················· | ···················· | ···················· |
| J | 8774 hCoV-19/England/ALDP-9533FD/2020\|EPI_ISL_554534\|20 | ···················· | ···················· | ···················· | ···················· | ···················· | ···················· |
| J | 8775 hCoV-19/England/ALDP-950C22/2020\|EPI_ISL_554535\|20 | ···················· | ···················· | ···················· | ···················· | ···················· | ···················· |
| J | 8776 hCoV-19/England/ALDP-953375/2020\|EPI_ISL_554536\|20 | ···················· | ·A·················· | ···················· | ···················· | ···················· | ···················· |
| J | 8777 hCoV-19/England/ALDP-951508/2020\|EPI_ISL_554539\|20 | ···················· | ···················· | ···················· | ···················· | ···················· | ···················· |
| J | 8778 hCoV-19/England/ALDP-95159F/2020\|EPI_ISL_554540\|20 | ···················· | ···················· | ···················· | ···················· | ···················· | ···················· |
| J | 8779 hCoV-19/England/ALDP-955786/2020\|EPI_ISL_554542\|20 | ···················· | ···················· | ···················· | ···················· | ···················· | ···················· |
| J | 8780 hCoV-19/England/ALDP-9532F0/2020\|EPI_ISL_554543\|20 | ···················· | ···················· | ···················· | ···················· | ···················· | ···················· |
| J | 8781 hCoV-19/England/ALDP-9533EE/2020\|EPI_ISL_554544\|20 | ···················· | ···················· | ···················· | ···················· | ···················· | ···················· |
| J | 8782 hCoV-19/England/ALDP-950C9B/2020\|EPI_ISL_554545\|20 | ···················· | ···················· | ···················· | ···················· | ···················· | ···················· |
| J | 8783 hCoV-19/England/ALDP-953454/2020\|EPI_ISL_554546\|20 | ···················· | ···················· | ···················· | ···················· | ···················· | ···················· |
| J | 8784 hCoV-19/England/ALDP-95318A/2020\|EPI_ISL_554547\|20 | ···················· | ···················· | ···················· | ···················· | ···················· | ···················· |
| J | 8785 hCoV-19/England/ALDP-951201/2020\|EPI_ISL_554549\|20 | ···················· | ···················· | ···················· | ···················· | ···················· | ···················· |
| J | 8786 hCoV-19/England/ALDP-955777/2020\|EPI_ISL_554550\|20 | ···················· | ···················· | ···················· | ···················· | ···················· | ···················· |
| J | 8787 hCoV-19/England/ALDP-95580B/2020\|EPI_ISL_554551\|20 | ···················· | ···················· | ···················· | ···················· | ···················· | ···················· |
| J | 8788 hCoV-19/England/ALDP-955416/2020\|EPI_ISL_554552\|20 | ···················· | ···················· | ···················· | ···················· | ···················· | ···················· |
| J | 8789 hCoV-19/England/ALDP-9531D5/2020\|EPI_ISL_554553\|20 | ···················· | ···················· | ···················· | ···················· | ···················· | ···················· |
| J | 8790 hCoV-19/England/ALDP-951210/2020\|EPI_ISL_554554\|20 | ···················· | ···················· | ···················· | ···················· | ···················· | ···················· |
| J | 8791 hCoV-19/England/ALDP-953481/2020\|EPI_ISL_554555\|20 | ···················· | ···················· | ···················· | ···················· | ···················· | ···················· |
| J | 8792 hCoV-19/England/ALDP-951641/2020\|EPI_ISL_554556\|20 | ···················· | ···················· | ···················· | ···················· | ···················· | ···················· |
| J | 8793 hCoV-19/England/ALDP-951429/2020\|EPI_ISL_554557\|20 | ···················· | ···················· | ···················· | ···················· | ···················· | ···················· |
| J | 8794 hCoV-19/England/ALDP-95085B/2020\|EPI_ISL_554558\|20 | ···················· | ···················· | ···················· | ···················· | ···················· | ···················· |

J 8795 hCoV-19/England/ALDP-95567A/2020|EPI_ISL_554560|20 ···················· ···················· ···················· ···················· ···················· ····················
J 8796 hCoV-19/England/ALDP-953C5C/2020|EPI_ISL_554561|20 ···················· ···················· ···················· ···················· ···················· ····················
J 8797 hCoV-19/England/ALDP-950B07/2020|EPI_ISL_554562|20 ···················· ···················· ···················· ···················· ···················· ····················
J 8798 hCoV-19/England/ALDP-950888/2020|EPI_ISL_554563|20 ···················· ···················· ···················· ···················· ···················· ····················
J 8799 hCoV-19/England/ALDP-95330C/2020|EPI_ISL_554564|20 ···················· ···················· ···················· ···················· ···················· ····················
J 8800 hCoV-19/Wales/ALDP-955856/2020|EPI_ISL_554565|2020 ···················· ···················· ···················· ···················· ···················· ····················
J 8801 hCoV-19/England/ALDP-950D2F/2020|EPI_ISL_554566|20 ···················· ···················· ···················· ···················· ···················· ····················
J 8802 hCoV-19/England/ALDP-94FF0F/2020|EPI_ISL_555810|20 ···················· ···················· ···················· ···················· ···················· ····················
J 8803 hCoV-19/England/ALDP-94B677/2020|EPI_ISL_555811|20 ···················· ···················· ···················· ···················· ···················· ····················
J 8804 hCoV-19/England/ALDP-94F8B9/2020|EPI_ISL_555812|20 ··C················· ···················· ···················· ···················· ···················· ····················
J 8805 hCoV-19/England/ALDP-94B783/2020|EPI_ISL_555814|20 ···················· ···················· ···················· ···················· ···················· ····················
J 8806 hCoV-19/England/ALDP-94F89B/2020|EPI_ISL_555816|20 ···················· ···················· ···················· ···················· ···················· ····················
J 8807 hCoV-19/England/ALDP-94A99D/2020|EPI_ISL_555817|20 ···················· ···················· ···················· ···················· ···················· ····················
J 8808 hCoV-19/England/ALDP-94B6B3/2020|EPI_ISL_555818|20 ···················· ···················· ···················· ···················· ···················· ····················
J 8809 hCoV-19/England/ALDP-94FF1E/2020|EPI_ISL_555820|20 ···················· ···················· ···················· ···················· ···················· ····················
J 8810 hCoV-19/England/ALDP-94BB4B/2020|EPI_ISL_555821|20 ···················· ···················· ···················· ···················· ···················· ····················
J 8811 hCoV-19/England/ALDP-94B3BC/2020|EPI_ISL_555823|20 ···················· ···················· ···················· ···················· ···················· ····················
J 8812 hCoV-19/England/ALDP-94B13A/2020|EPI_ISL_555824|20 ···················· ···················· ···················· ···················· ···················· ····················
J 8813 hCoV-19/England/ALDP-94AA03/2020|EPI_ISL_555825|20 ···················· ···················· ···················· ···················· ···················· ····················
J 8814 hCoV-19/England/ALDP-94B941/2020|EPI_ISL_555827|20 ···················· ···················· ···················· ···················· ···················· ····················
J 8815 hCoV-19/England/ALDP-94A9F7/2020|EPI_ISL_555830|20 ···················· ···················· ···················· ···················· ···················· ····················
J 8816 hCoV-19/England/ALDP-94F8E6/2020|EPI_ISL_555831|20 ···················· ···················· ···················· ···················· ···················· ····················
J 8817 hCoV-19/England/ALDP-94C843/2020|EPI_ISL_555832|20 ···················· ···················· ···················· ···················· ···················· ····················
J 8818 hCoV-19/England/ALDP-94D0E0/2020|EPI_ISL_555834|20 ···················· ···················· ···················· ···················· ···················· ····················
J 8819 hCoV-19/England/ALDP-94CEBA/2020|EPI_ISL_555836|20 ···················· ···················· ···················· ···················· ···················· ····················
J 8820 hCoV-19/England/ALDP-94CE23/2020|EPI_ISL_555837|20 ···················· ···················· ···················· ···················· ···················· ····················
J 8821 hCoV-19/England/ALDP-94F251/2020|EPI_ISL_555838|20 ···················· ···················· ···················· ···················· ···················· ····················
J 8822 hCoV-19/England/ALDP-94C8DA/2020|EPI_ISL_555839|20 ···················· ···················· ···················· ···················· ···················· ····················
J 8823 hCoV-19/England/ALDP-94CA89/2020|EPI_ISL_555840|20 ···················· ···················· ···················· ···················· ···················· ····················
J 8824 hCoV-19/England/ALDP-94CA5C/2020|EPI_ISL_555841|20 ···················· ···················· ···················· ···················· ···················· ····················
J 8825 hCoV-19/England/ALDP-94F233/2020|EPI_ISL_555842|20 ···················· ···················· ···················· ···················· ···················· ····················
J 8826 hCoV-19/England/ALDP-94C9F5/2020|EPI_ISL_555845|20 ···················· ···················· ···················· ···················· ···················· ····················
J 8827 hCoV-19/England/ALDP-94F41F/2020|EPI_ISL_555846|20 ···················· ···················· ···················· ···················· ···················· ····················
J 8828 hCoV-19/England/ALDP-94CEAB/2020|EPI_ISL_555847|20 ···················· ···················· ···················· ···················· ···················· ····················
J 8829 hCoV-19/England/ALDP-94C6FE/2020|EPI_ISL_555849|20 ···················· ···················· ···················· ···················· ···················· ····················
J 8830 hCoV-19/England/ALDP-94C904/2020|EPI_ISL_555850|20 ···················· ···················· ···················· ···················· ···················G ···················· 1
J 8831 hCoV-19/England/ALDP-94CF5D/2020|EPI_ISL_555851|20 ···················· ···················· ···················· ···················· ···················· ····················
J 8832 hCoV-19/England/ALDP-94C7FB/2020|EPI_ISL_555852|20 ···················· ···················· ···················· ···················· ···················· ····················
J 8833 hCoV-19/England/ALDP-94C676/2020|EPI_ISL_555854|20 ···················· ···················· ···················· ···················· ···················· ····················
J 8834 hCoV-19/England/ALDP-94F260/2020|EPI_ISL_555855|20 ···················· ···················· ···················· ···················· ···················· ····················
J 8835 hCoV-19/England/ALDP-94CBD1/2020|EPI_ISL_555856|20 ···················· ···················· ···················· ···················· ···················· ····················
J 8836 hCoV-19/England/ALDP-94CD08/2020|EPI_ISL_555857|20 ···················· ···················· ···················· ···················· ···················· ····················
J 8837 hCoV-19/England/ALDP-94CC0B/2020|EPI_ISL_555858|20 ···················· ···················· ···················· ···················· ···················· ····················
J 8838 hCoV-19/England/ALDP-94F29D/2020|EPI_ISL_555859|20 ···················· ···················· ···················· ···················· ···················· ····················
J 8839 hCoV-19/Wales/ALDP-94C773/2020|EPI_ISL_555860|2020 ···················· ···················· ···················· ···················· ···················· ····················
J 8840 hCoV-19/England/ALDP-94CAF2/2020|EPI_ISL_555861|20 ···················· ···················· ···················· ···················· ···················· ····················
J 8841 hCoV-19/England/ALDP-94CBC2/2020|EPI_ISL_555862|20 ···················· ···················· ···················· ···················· ···················· ····················
J 8842 hCoV-19/England/ALDP-94C95F/2020|EPI_ISL_555863|20 ···················· ···················· ···················· ···················· ···················· ····················
J 8843 hCoV-19/England/CAMC-949688/2020|EPI_ISL_555864|20 ···················· ···················· ···················· ···················· ···················· ····················
J 8844 hCoV-19/England/ALDP-94C649/2020|EPI_ISL_555865|20 ···················· ···················· ···················· ···················· ···················· ····················
J 8845 hCoV-19/England/ALDP-94C3E8/2020|EPI_ISL_555866|20 ···················· ···················· ···················· ···················· ···················· ····················
J 8846 hCoV-19/England/ALDP-94C685/2020|EPI_ISL_555867|20 ···················· ···················· ···················· ···················· ···················· ····················
J 8847 hCoV-19/England/CAMC-948883/2020|EPI_ISL_555868|20 ···················· ···················· ···················· ···················· ···················· ····················
J 8848 hCoV-19/England/ALDP-94F35E/2020|EPI_ISL_555869|20 ···················· ···················· ···················· ···················· ···················· ····················
J 8849 hCoV-19/England/ALDP-94F646/2020|EPI_ISL_555870|20 ···················· ···················· ···················· ···················· ···················· ····················
J 8850 hCoV-19/England/ALDP-94C54C/2020|EPI_ISL_555871|20 ···················· ···················· ···················· ···················· ···················· ····················
J 8851 hCoV-19/England/ALDP-94CE05/2020|EPI_ISL_555872|20 ···················· ···················· ···················· ···················· ···················· ····················
J 8852 hCoV-19/Wales/ALDP-94F3A9/2020|EPI_ISL_555873|2020 ···················· ···················· ···················· ···················· ···················· ····················
J 8853 hCoV-19/England/ALDP-94C5C4/2020|EPI_ISL_555874|20 ···················· ···················· ···················· ···················· ···················· ····················
J 8854 hCoV-19/England/CAMC-948CF0/2020|EPI_ISL_555875|20 ···················· ···················· ···················· ···················· ···················· ····················
J 8855 hCoV-19/England/ALDP-94C588/2020|EPI_ISL_555876|20 ···················· ···················· ···················· ···················· ···················· ····················
J 8856 hCoV-19/England/ALDP-94C728/2020|EPI_ISL_555878|20 ···················· ···················· ···················· ···················· ···················· ····················
J 8857 hCoV-19/England/ALDP-94D03B/2020|EPI_ISL_555879|20 ···················· ···················· ···················· ···················· ···················· ····················
J 8858 hCoV-19/England/ALDP-94CA98/2020|EPI_ISL_555880|20 ···················· ···················· ···················· ···················· ···················· ····················
J 8859 hCoV-19/England/ALDP-94F60A/2020|EPI_ISL_555881|20 ···················· ·A·················· ···················· ···················· ···················· ····················
J 8860 hCoV-19/England/ALDP-94CE7E/2020|EPI_ISL_555882|20 ···················· ···················· ···················· ···················· ···················· ····················
J 8861 hCoV-19/England/ALDP-94CDAE/2020|EPI_ISL_555883|20 ···················· ···A················ ···················· ···················· ···················· ····················
J 8862 hCoV-19/Wales/ALDP-94F585/2020|EPI_ISL_555884|2020 ···················· ···················· ···················· ···················· ···················· ····················
J 8863 hCoV-19/England/ALDP-94C421/2020|EPI_ISL_555885|20 ···················· ···················· ···················· ···················· ···················· ····················
J 8864 hCoV-19/England/ALDP-94C4B8/2020|EPI_ISL_555886|20 ···················· ···················· ···················· ···················· ···················· ····················
J 8865 hCoV-19/Wales/ALDP-94C737/2020|EPI_ISL_555888|2020 ···················· ···················· ···················· ···················· ···················· ····················
J 8866 hCoV-19/England/ALDP-94D02C/2020|EPI_ISL_555889|20 ···················· ···················· ···················· ···················· ···················· ····················
J 8867 hCoV-19/England/ALDP-94CD44/2020|EPI_ISL_555890|20 ···················· ···················· ···················· ···················· ···················· ····················
J 8868 hCoV-19/England/ALDP-94C70A/2020|EPI_ISL_555891|20 ···················· ·A·················· ···················· ···················· ···················· ····················
J 8869 hCoV-19/England/ALDP-94F093/2020|EPI_ISL_555892|20 ···················· ···················· ···················· ···················· ···················· ····················
J 8870 hCoV-19/England/ALDP-94CA3E/2020|EPI_ISL_555893|20 ···················· ···················· ···················· ···················· ···················· ····················
J 8871 hCoV-19/England/ALDP-94F4E2/2020|EPI_ISL_555894|20 ···················· ···················· ···················· ···················· ···················· ····················
J 8872 hCoV-19/Wales/ALDP-94F567/2020|EPI_ISL_555895|2020 ···················· ···················· ···················· ···················· ···················· ····················
J 8873 hCoV-19/England/ALDP-94CE6F/2020|EPI_ISL_555896|20 ···················· ···················· ···················· ···················· ···················· ····················
J 8874 hCoV-19/England/ALDP-94C834/2020|EPI_ISL_555897|20 ···················· ···················· ···················· ···················· ···················· ····················

| | | | | | | | | |
|---|---|---|---|---|---|---|---|---|
| J | 8875 | hCoV-19/England/ALDP-68C2D3/2020\|EPI_ISL_558010\|20 | ···················· | ···················· | ····················· | ···················· | ···················· | ···················· |
| J | 8876 | hCoV-19/England/ALDP-691174/2020\|EPI_ISL_558011\|20 | ···················· | ···················· | ····················· | ···················· | ···················· | ···················· |
| J | 8877 | hCoV-19/England/ALDP-6461AB/2020\|EPI_ISL_558012\|20 | ···················· | ······A············· | ····················· | ···················· | ···················· | ···················· |
| J | 8878 | hCoV-19/England/ALDP-68C376/2020\|EPI_ISL_558013\|20 | ···················· | ···················· | ····················· | ···················· | ···················· | ···················· |
| J | 8879 | hCoV-19/England/ALDP-66DCCB/2020\|EPI_ISL_558014\|20 | ···················· | ···················· | ····················· | ···················· | ···················· | ···················· |
| J | 8880 | hCoV-19/England/ALDP-644B72/2020\|EPI_ISL_558015\|20 | ···················· | ···················· | ····················· | ···················· | ···················· | ···················· |
| J | 8881 | hCoV-19/England/ALDP-65C98C/2020\|EPI_ISL_558018\|20 | ···················· | ···················· | ····················· | ···················· | ···················· | ···················· |
| J | 8882 | hCoV-19/England/ALDP-6598DD/2020\|EPI_ISL_558019\|20 | ···················· | ···················· | ····················· | ···················· | ···················· | ···················· |
| J | 8883 | hCoV-19/England/MILK-64F4B8/2020\|EPI_ISL_558021\|20 | ···················· | ···················· | ····················· | ···················· | ···················· | ···················· |
| J | 8884 | hCoV-19/England/ALDP-646439/2020\|EPI_ISL_558022\|20 | ···················· | ···················· | ····················· | ···················· | ···················· | ···················· |
| J | 8885 | hCoV-19/England/ALDP-6440BF/2020\|EPI_ISL_558025\|20 | ···················· | ···················· | ····················· | ···················· | ···················· | ···················· |
| J | 8886 | hCoV-19/England/MILK-66CDAB/2020\|EPI_ISL_558026\|20 | ···················· | ···················· | ····················· | ···················· | ···················· | ···················· |
| J | 8887 | hCoV-19/England/ALDP-658FAC/2020\|EPI_ISL_558027\|20 | ···················· | ···················· | ····················· | ···················· | ···················· | ···················· |
| J | 8888 | hCoV-19/England/MILK-654EC1/2020\|EPI_ISL_558028\|20 | ···················· | ···················· | ····················· | ···················· | ···················· | ···················· |
| J | 8889 | hCoV-19/England/ALDP-66C4E2/2020\|EPI_ISL_558029\|20 | ···················· | ···················· | ····················· | ···················· | ···················· | ···················· |
| J | 8890 | hCoV-19/England/ALDP-68EE37/2020\|EPI_ISL_558032\|20 | ···················· | ···················· | ····················· | ···················· | ···················· | ···················· |
| J | 8891 | hCoV-19/England/ALDP-6511CB/2020\|EPI_ISL_558033\|20 | ···················· | ···················· | ····················· | ···················· | ···················· | ···················· |
| J | 8892 | hCoV-19/England/ALDP-6763E8/2020\|EPI_ISL_558034\|20 | ···················· | ···················· | ····················· | ···················· | ···················· | ···················· |
| J | 8893 | hCoV-19/England/ALDP-66C840/2020\|EPI_ISL_558036\|20 | ···················· | ···················· | ····················· | ···················· | ···················· | ···················· |
| J | 8894 | hCoV-19/England/MILK-65409E/2020\|EPI_ISL_558037\|20 | ···················· | ···················· | ····················· | ···················· | ···················· | ···················· |
| J | 8895 | hCoV-19/England/ALDP-670050/2020\|EPI_ISL_558040\|20 | ···················· | ···················· | ····················· | ···················· | ···················· | ···················· |
| J | 8896 | hCoV-19/England/MILK-64FFB7/2020\|EPI_ISL_558041\|20 | ···················· | ·A·················· | ····················· | ···················· | ···················· | ···A················ |
| J | 8897 | hCoV-19/England/ALDP-651AEE/2020\|EPI_ISL_558042\|20 | ···················· | ···················· | ····················· | ···················· | ···················· | ···················· |
| J | 8898 | hCoV-19/England/ALDP-6901C0/2020\|EPI_ISL_558043\|20 | ···················· | ···················· | ····················· | ···················· | ···················· | ···················· |
| J | 8899 | hCoV-19/England/ALDP-6511BC/2020\|EPI_ISL_558044\|20 | ···················· | ···················· | ····················· | ···················· | ···················· | ···················· |
| J | 8900 | hCoV-19/England/ALDP-6471E6/2020\|EPI_ISL_558045\|20 | ···················· | ···················· | ····················· | ···················· | ···················· | ···················· |
| J | 8901 | hCoV-19/England/ALDP-649D77/2020\|EPI_ISL_558046\|20 | ···················· | ···················· | ····················· | ···················· | ···················· | ···················· |
| J | 8902 | hCoV-19/England/ALDP-65859B/2020\|EPI_ISL_558047\|20 | ···················· | ···················· | ····················· | ···················· | ···················· | ···················· |
| J | 8903 | hCoV-19/England/ALDP-65858C/2020\|EPI_ISL_558048\|20 | ···················· | ···················· | ····················· | ···················· | ···················· | ···················· |
| J | 8904 | hCoV-19/England/ALDP-64AC5B/2020\|EPI_ISL_558049\|20 | ···················· | ···················· | ····················· | ···················· | ···················· | ···················· |
| J | 8905 | hCoV-19/England/ALDP-68C19A/2020\|EPI_ISL_558060\|20 | ···················· | ···················· | ····················· | ···················· | ···················· | ···················· |
| J | 8906 | hCoV-19/England/ALDP-68DBB9/2020\|EPI_ISL_558061\|20 | ···················· | ···················· | ····················· | ···················· | ···················· | ···················· |
| J | 8907 | hCoV-19/England/ALDP-6485BC/2020\|EPI_ISL_558062\|20 | ···················· | ·A·················· | ····················· | ···················· | ···················· | ···················· |
| J | 8908 | hCoV-19/England/ALDP-645F75/2020\|EPI_ISL_558063\|20 | ···················· | ······A············· | ····················· | ···················· | ···················· | ···················· |
| J | 8909 | hCoV-19/England/ALDP-6913BA/2020\|EPI_ISL_558090\|20 | ···················· | ···················· | ····················· | ···················· | ···················· | ···················· |
| J | 8910 | hCoV-19/England/ALDP-655FBE/2020\|EPI_ISL_558091\|20 | ···················· | ···················· | ····················· | ···················· | ···················· | ···················· |
| J | 8911 | hCoV-19/England/ALDP-691CFB/2020\|EPI_ISL_558092\|20 | ···················· | ···················· | ····················· | ···················· | ···················· | ···················· |
| J | 8912 | hCoV-19/England/ALDP-68C2E2/2020\|EPI_ISL_558093\|20 | ···················· | ···················· | ····················· | ···················· | ···················· | ···················· |
| J | 8913 | hCoV-19/England/ALDP-68F3CE/2020\|EPI_ISL_558094\|20 | ···················· | ···················· | ····················· | ···················· | ···················· | ···················· |
| J | 8914 | hCoV-19/England/ALDP-66F661/2020\|EPI_ISL_558095\|20 | ···················· | ·A·················· | ····················· | ···················· | ···················· | ···················· |
| J | 8915 | hCoV-19/England/MILK-66C93E/2020\|EPI_ISL_558096\|20 | ···················· | ···················· | ····················· | ···················· | ···················· | ···················· |
| J | 8916 | hCoV-19/England/ALDP-6570D7/2020\|EPI_ISL_558097\|20 | ···················· | ···················· | ····················· | ···················· | ···················· | ···················· |
| J | 8917 | hCoV-19/England/ALDP-650AB2/2020\|EPI_ISL_558098\|20 | ···················· | ···················· | ····················· | ···················· | ···················· | ···················· |
| J | 8918 | hCoV-19/England/ALDP-64642A/2020\|EPI_ISL_558099\|20 | ···················· | ···················· | ····················· | ···················· | ···················· | ···················· |
| J | 8919 | hCoV-19/England/ALDP-49B631/2020\|EPI_ISL_559340\|20 | ···················· | ···················· | ····················· | ···················· | ···················· | ···················· |
| J | 8920 | hCoV-19/England/ALDP-4AD362/2020\|EPI_ISL_559341\|20 | ···················· | ···················· | ····················· | ···················· | ···················· | ···················· |
| J | 8921 | hCoV-19/England/ALDP-4AED45/2020\|EPI_ISL_559342\|20 | ···················· | ···················· | ····················· | ···················· | ···················· | ···················· |
| J | 8922 | hCoV-19/England/ALDP-4AEBF0/2020\|EPI_ISL_559343\|20 | ···················· | ···················· | ····················· | ···················· | ···················· | ···················· |
| J | 8923 | hCoV-19/England/ALDP-496B82/2020\|EPI_ISL_559344\|20 | ···················· | ···················· | ····················· | ···················· | ···················· | ···················· |
| J | 8924 | hCoV-19/England/ALDP-4971F8/2020\|EPI_ISL_559345\|20 | ···················· | ···················· | ····················· | ···················· | ···················· | ···················· |
| J | 8925 | hCoV-19/England/ALDP-4AEB1E/2020\|EPI_ISL_559346\|20 | ···················· | ···················· | ····················· | ···················· | ···················· | ···················· |
| J | 8926 | hCoV-19/England/ALDP-49B9B0/2020\|EPI_ISL_559347\|20 | ···················· | ···················· | ····················· | ···················· | ···················· | ···················· |
| J | 8927 | hCoV-19/England/ALDP-4AE99C/2020\|EPI_ISL_559348\|20 | ···················· | ···················· | ····················· | ···················· | ···················· | ···················· |
| J | 8928 | hCoV-19/England/ALDP-497538/2020\|EPI_ISL_559349\|20 | ···················· | ···················· | ····················· | ···················· | ···················· | ···················· |
| J | 8929 | hCoV-19/England/ALDP-496D40/2020\|EPI_ISL_559351\|20 | ···················· | ···················· | ····················· | ···················· | ···················· | ···················· |
| J | 8930 | hCoV-19/England/MILK-4A036F/2020\|EPI_ISL_559352\|20 | ···················· | ···················· | ····················· | ···················· | ···················· | ···················· |
| J | 8931 | hCoV-19/England/MILK-4A1234/2020\|EPI_ISL_559354\|20 | ···················· | ···················· | ····················· | ···················· | ···················· | ···················· |
| J | 8932 | hCoV-19/England/MILK-4A045D/2020\|EPI_ISL_559355\|20 | ···················· | ···················· | ····················· | ···················· | ···················· | ···················· |
| J | 8933 | hCoV-19/England/ALDP-49AC6D/2020\|EPI_ISL_559356\|20 | ···················· | ···················· | ····················· | ···················· | ···················· | ···················· |
| J | 8934 | hCoV-19/England/ALDP-4AD5A8/2020\|EPI_ISL_559358\|20 | ···················· | ···················· | ····················· | ···················· | ···················· | ···················· |
| J | 8935 | hCoV-19/England/ALDP-49692E/2020\|EPI_ISL_559359\|20 | ···················· | ···················· | ····················· | ···················· | ···················· | ···················· |
| J | 8936 | hCoV-19/England/ALDP-49B74D/2020\|EPI_ISL_559360\|20 | ···················· | ···················· | ····················· | ···················· | ···················· | ···················· |
| J | 8937 | hCoV-19/England/ALDP-497565/2020\|EPI_ISL_559361\|20 | ···················· | ···················· | ····················· | ···················· | ···················· | ···················· |
| J | 8938 | hCoV-19/England/ALDP-4AEE24/2020\|EPI_ISL_559362\|20 | ···················· | ···················· | ····················· | ···················· | ···················· | ···················· |
| J | 8939 | hCoV-19/England/ALDP-4AEA20/2020\|EPI_ISL_559363\|20 | ···················· | ···················· | ····················· | ···················· | ···················· | ···················· |
| J | 8940 | hCoV-19/England/ALDP-4AEE15/2020\|EPI_ISL_559365\|20 | ···················· | ···················· | ····················· | ···················· | ···················· | ···················· |
| J | 8941 | hCoV-19/England/ALDP-49B200/2020\|EPI_ISL_559366\|20 | ···················· | ···················· | ····················· | ···················· | ···················· | ···················· |
| J | 8942 | hCoV-19/England/ALDP-4AD423/2020\|EPI_ISL_559367\|20 | ···················· | ···················· | ····················· | ···················· | ···················· | ···················· |
| J | 8943 | hCoV-19/England/MILK-4A0077/2020\|EPI_ISL_559368\|20 | ···················· | ···················· | ····················· | ···················· | ···················· | ···················· |
| J | 8944 | hCoV-19/England/ALDP-49B0E8/2020\|EPI_ISL_559369\|20 | ···················· | ···················· | ····················· | ···················· | ···················· | ···················· |
| J | 8945 | hCoV-19/England/ALDP-4AB258/2020\|EPI_ISL_559370\|20 | ···················· | ···················· | ····················· | ···················· | ···················· | ···················· |
| J | 8946 | hCoV-19/England/ALDP-49978B/2020\|EPI_ISL_559371\|20 | ···················· | ···················· | ····················· | ···················· | ···················· | ···················· |
| J | 8947 | hCoV-19/England/ALDP-499633/2020\|EPI_ISL_559372\|20 | ···················· | ···················· | ····················· | ···················· | ···················· | ···················· |
| J | 8948 | hCoV-19/England/ALDP-4A229D/2020\|EPI_ISL_559375\|20 | ···················· | ···················· | ····················· | ···················· | ···················· | ···················· |
| J | 8949 | hCoV-19/England/ALDP-4982D6/2020\|EPI_ISL_559376\|20 | ···················· | ···················· | ····················· | ···················· | ···················· | ···················· |
| J | 8950 | hCoV-19/England/ALDP-4983D3/2020\|EPI_ISL_559377\|20 | ···················· | ···················· | ····················· | ···················· | ···················· | ···················· |
| J | 8951 | hCoV-19/England/ALDP-4A1DBB/2020\|EPI_ISL_559378\|20 | ···················· | ···················· | ····················· | ···················· | ···················· | ···················· |
| J | 8952 | hCoV-19/England/ALDP-498607/2020\|EPI_ISL_559379\|20 | ···················· | ···················· | ····················· | ···················· | ···················· | ···················· |
| J | 8953 | hCoV-19/England/ALDP-497CBB/2020\|EPI_ISL_559392\|20 | ···················· | ···················· | ····················· | ···················· | ···················· | ···················· |
| J | 8954 | hCoV-19/England/ALDP-498A0B/2020\|EPI_ISL_559393\|20 | ···················· | ···················· | ·········T··········· | ···················· | ···················· | ···················· |

| | | | | | | | | |
|---|---|---|---|---|---|---|---|---|
| J | 8955 | hCoV-19/England/ALDP-49A30E/2020\|EPI_ISL_559394\|20 | ···················· | ···················· | ····················· | ···················· | ···················· | ···················· |
| J | 8956 | hCoV-19/India/UT-IMT-AT52/2020\|EPI_ISL_560320\|2020 | ···················· | ·A·················· | ····················· | ···················· | ···················· | ···A················ |
| J | 8957 | hCoV-19/India/UT-IMT-BB45/2020\|EPI_ISL_560321\|2020 | ···················· | ·A·················· | ····················· | ···················· | ···················· | ···A················ |
| J | 8958 | hCoV-19/India/HR-IMT-BZ115/2020\|EPI_ISL_560322\|202 | ···················· | ···················· | ····················· | ···················· | ···················· | ···················· |
| J | 8959 | hCoV-19/India/HR-IMT-BZ119/2020\|EPI_ISL_560323\|202 | ···················· | ···················· | ····················· | ···················· | ···················· | ···················· |
| J | 8960 | hCoV-19/India/HR-IMT-CB117/2020\|EPI_ISL_560324\|202 | ···················· | ·A·················· | ····················· | ···················· | ···················· | ···A················ |
| J | 8961 | hCoV-19/England/ALDP-94B53E/2020\|EPI_ISL_555800\|20 | ···················· | ···················· | ····················· | ···················· | ···················· | ···················· |
| J | 8962 | hCoV-19/England/ALDP-94FAB3/2020\|EPI_ISL_555801\|20 | ···················· | ···················· | ····················· | ···················· | ···················· | ···················· |
| J | 8963 | hCoV-19/England/ALDP-94BB69/2020\|EPI_ISL_555802\|20 | ···················· | ···················· | ····················· | ···················· | ···················· | ···················· |
| J | 8964 | hCoV-19/England/ALDP-94F8C8/2020\|EPI_ISL_555803\|20 | ···················· | ···················· | ····················· | ···················· | ···················· | ···················· |
| J | 8965 | hCoV-19/England/ALDP-94F743/2020\|EPI_ISL_555804\|20 | ···················· | ···················· | ····················· | ···················· | ···················· | ···················· |
| J | 8966 | hCoV-19/England/ALDP-94B39E/2020\|EPI_ISL_555805\|20 | ···················· | ···················· | ····················· | ···················· | ···················· | ···················· |
| J | 8967 | hCoV-19/England/ALDP-94B38F/2020\|EPI_ISL_555806\|20 | ···················· | ···R················ | ····················· | ···················· | ···················· | ···················· |
| J | 8968 | hCoV-19/England/ALDP-94F7DA/2020\|EPI_ISL_555808\|20 | ···················· | ···················· | ····················· | ···················· | ···················· | ···················· |
| J | 8969 | hCoV-19/England/ALDP-66DEF2/2020\|EPI_ISL_558050\|20 | ···················· | ···················· | ····················· | ···················· | ···················· | ···················· |
| J | 8970 | hCoV-19/England/ALDP-68F4BC/2020\|EPI_ISL_558051\|20 | ···················· | ···················· | ····················· | ···················· | ···················· | ···················· |
| J | 8971 | hCoV-19/England/ALDP-673BD4/2020\|EPI_ISL_558053\|20 | ···················· | ···················· | ····················· | ···················· | ···················· | ···················· |
| J | 8972 | hCoV-19/England/ALDP-68FE81/2020\|EPI_ISL_558054\|20 | ···················· | ···················· | ····················· | ···················· | ···················· | ···················· |
| J | 8973 | hCoV-19/England/ALDP-64C22A/2020\|EPI_ISL_558057\|20 | ···················· | ···················· | ····················· | ···················· | ···················· | ···················· |
| J | 8974 | hCoV-19/England/MILK-671CA6/2020\|EPI_ISL_558058\|20 | ···················· | ···················· | ····················· | ···················· | ···················· | ···················· |
| J | 8975 | hCoV-19/England/ALDP-65C2AF/2020\|EPI_ISL_558059\|20 | ···················· | ···················· | ····················· | ···················· | ···················· | ···················· |
| J | 8976 | hCoV-19/England/ALDP-673C86/2020\|EPI_ISL_558066\|20 | ···················· | ···················· | ····················· | ···················· | ···················· | ···················· |
| J | 8977 | hCoV-19/England/ALDP-64B48F/2020\|EPI_ISL_558067\|20 | ···················· | ···················· | ····················· | ···················· | ···················· | ···················· |
| J | 8978 | hCoV-19/England/MILK-674F22/2020\|EPI_ISL_558068\|20 | ···················· | ···················· | ····················· | ···················· | ···················· | ···················· |
| J | 8979 | hCoV-19/England/ALDP-66E617/2020\|EPI_ISL_558070\|20 | ···················· | ···················· | ····················· | ···················· | ···················· | ···················· |
| J | 8980 | hCoV-19/England/ALDP-645670/2020\|EPI_ISL_558072\|20 | ···················· | ···················· | ····················· | ···················· | ···················· | ···················· |
| J | 8981 | hCoV-19/England/ALDP-68E040/2020\|EPI_ISL_558074\|20 | ···················· | ···················· | ····················· | ···················· | ···················· | ···················· |
| J | 8982 | hCoV-19/England/ALDP-673B7A/2020\|EPI_ISL_558075\|20 | ···················· | ···················· | ····················· | ···················· | ···················· | ···················· |
| J | 8983 | hCoV-19/England/ALDP-649463/2020\|EPI_ISL_558076\|20 | ···················· | ···················· | ····················· | ···················· | ···················· | ···················· |
| J | 8984 | hCoV-19/England/ALDP-676236/2020\|EPI_ISL_558077\|20 | ···················· | ···················· | ····················· | ···················· | ···················· | ···················· |
| J | 8985 | hCoV-19/England/ALDP-656DFF/2020\|EPI_ISL_558078\|20 | ···················· | ···················· | ····················· | ···················· | ···················· | ···················· |
| J | 8986 | hCoV-19/England/ALDP-6761B1/2020\|EPI_ISL_558080\|20 | ···················· | ···················· | ····················· | ···················· | ···················· | ···················· |
| J | 8987 | hCoV-19/England/ALDP-65615D/2020\|EPI_ISL_558081\|20 | ···················· | ···················· | ····················· | ···················· | ···················· | ···················· |
| J | 8988 | hCoV-19/England/ALDP-6530CC/2020\|EPI_ISL_558082\|20 | ···················· | ···················· | ····················· | ···················· | ···················· | ···················· |
| J | 8989 | hCoV-19/England/ALDP-64D432/2020\|EPI_ISL_558084\|20 | ···················· | ···················· | ····················· | ···················· | ···················· | ···················· |
| J | 8990 | hCoV-19/England/ALDP-658C87/2020\|EPI_ISL_558085\|20 | ···················· | ···················· | ····················· | ···················· | ···················· | ···················· |
| J | 8991 | hCoV-19/England/ALDP-671A24/2020\|EPI_ISL_558086\|20 | ···················· | ···················· | ····················· | ···················· | ···················· | ···················· |
| J | 8992 | hCoV-19/England/ALDP-64DEAD/2020\|EPI_ISL_558088\|20 | ···················· | ···················· | ····················· | ···················· | ···················· | ···················· |
| J | 8993 | hCoV-19/England/ALDP-49A41A/2020\|EPI_ISL_559380\|20 | ···················· | ···················· | ····················· | ···················· | ···················· | ···················· |
| J | 8994 | hCoV-19/England/ALDP-4997C7/2020\|EPI_ISL_559381\|20 | ···················· | ···················· | ····················· | ···················· | ···················· | ···················· |
| J | 8995 | hCoV-19/England/ALDP-4A21FA/2020\|EPI_ISL_559382\|20 | ···················· | ···················· | ····················· | ···················· | ···················· | ···················· |
| J | 8996 | hCoV-19/England/ALDP-49A605/2020\|EPI_ISL_559383\|20 | ···················· | ···················· | ····················· | ···················· | ···················· | ···················· |
| J | 8997 | hCoV-19/England/ALDP-499624/2020\|EPI_ISL_559384\|20 | ···················· | ···················· | ····················· | ···················· | ···················· | ···················· |
| J | 8998 | hCoV-19/England/ALDP-4990AE/2020\|EPI_ISL_559385\|20 | ···················· | ···················· | ····················· | ···················· | ···················· | ···················· |
| J | 8999 | hCoV-19/England/ALDP-49A456/2020\|EPI_ISL_559386\|20 | ···················· | ···················· | ····················· | ···················· | ···················· | ···················· |
| J | 9000 | hCoV-19/England/ALDP-4A279E/2020\|EPI_ISL_559388\|20 | ···················· | ···················· | ····················· | ···················· | ···················· | ···················· |
| J | 9001 | hCoV-19/England/ALDP-49A474/2020\|EPI_ISL_559395\|20 | ···················· | ···················· | ····················· | ···················· | ···················· | ···················· |
| J | 9002 | hCoV-19/England/ALDP-49942A/2020\|EPI_ISL_559396\|20 | ···················· | ···················· | ····················· | ···················· | ···················· | ···················· |
| J | 9003 | hCoV-19/England/ALDP-4A235E/2020\|EPI_ISL_559397\|20 | ···················· | ···················· | ····················· | ···················· | ···················· | ···················· |
| J | 9004 | hCoV-19/England/ALDP-499AFB/2020\|EPI_ISL_559398\|20 | ···················· | ···················· | ····················· | ···················· | ···················· | ···················· |
| J | 9005 | hCoV-19/England/ALDP-4A2A77/2020\|EPI_ISL_559399\|20 | ···················· | ···················· | ····················· | ···················· | ···················· | ···················· |
| K | 1 | hCoV-19/Australia/VIC11165/2020\|EPI_ISL_563279\|202 | ···················· | ···················· | ····················· | ···················· | ···················· | ···················· |
| K | 2 | hCoV-19/Australia/VIC11168/2020\|EPI_ISL_563280\|202 | ···················· | ···················· | ····················· | ···················· | ···················· | ···················· |
| K | 3 | hCoV-19/Australia/VIC11169/2020\|EPI_ISL_563281\|202 | ···················· | ···················· | ····················· | ···················· | ···················· | ···················· |
| K | 4 | hCoV-19/Australia/VIC11174/2020\|EPI_ISL_563282\|202 | ···················· | ···················· | ····················· | ···················· | ···················· | ···················· |
| K | 5 | hCoV-19/Australia/VIC11175/2020\|EPI_ISL_563283\|202 | ···················· | ···················· | ····················· | ···················· | ···················· | ···················· |
| K | 6 | hCoV-19/Australia/VIC11176/2020\|EPI_ISL_563284\|202 | ···················· | ···················· | ····················· | ···················· | ···················· | ···················· |
| K | 7 | hCoV-19/Australia/VIC11178/2020\|EPI_ISL_563285\|202 | ···················· | ···················· | ····················· | ···················· | ···················· | ···················· |
| K | 8 | hCoV-19/Australia/VIC11181/2020\|EPI_ISL_563286\|202 | ···················· | ···················· | ····················· | ···················· | ···················· | ···················· |
| K | 9 | hCoV-19/Australia/VIC11183/2020\|EPI_ISL_563287\|202 | ···················· | ···················· | ····················· | ···················· | ···················· | ···················· |
| K | 10 | hCoV-19/Australia/VIC11186/2020\|EPI_ISL_563288\|202 | ···················· | ···················· | ····················· | ···················· | ···················· | ···················· |
| K | 11 | hCoV-19/Australia/VIC11188/2020\|EPI_ISL_563289\|202 | ···················· | ···················· | ····················· | ···················· | ···················· | ···················· |
| K | 12 | hCoV-19/Australia/VIC11191/2020\|EPI_ISL_563290\|202 | ···················· | ···················· | ····················· | ···················· | ···················· | ···················· |
| K | 13 | hCoV-19/Australia/VIC11193/2020\|EPI_ISL_563291\|202 | ···················· | ···················· | ····················· | ···················· | ···················· | ···················· |
| K | 14 | hCoV-19/Australia/VIC11195/2020\|EPI_ISL_563292\|202 | ···················· | ···················· | ····················· | ···················· | ···················· | ···················· |
| K | 15 | hCoV-19/Australia/VIC11197/2020\|EPI_ISL_563293\|202 | ···················· | ···················· | ····················· | ···················· | ···················· | ···················· |
| K | 16 | hCoV-19/Australia/VIC11198/2020\|EPI_ISL_563294\|202 | ···················· | ···················· | ····················· | ···················· | ···················· | ···················· |
| K | 17 | hCoV-19/Australia/VIC11202/2020\|EPI_ISL_563295\|202 | ···················· | ···················· | ····················· | ···················· | ···················· | ···················· |
| K | 18 | hCoV-19/Australia/VIC11203/2020\|EPI_ISL_563296\|202 | ···················· | ···················· | ····················· | ···················· | ···················· | ···················· |
| K | 19 | hCoV-19/Australia/VIC11207/2020\|EPI_ISL_563297\|202 | ···················· | ···················· | ····················· | ···················· | ···················· | ···················· |
| K | 20 | hCoV-19/Australia/VIC11211/2020\|EPI_ISL_563298\|202 | ···················· | ···················· | ····················· | ···················· | ···················· | ···················· |
| K | 21 | hCoV-19/Australia/VIC11214/2020\|EPI_ISL_563299\|202 | ···················· | ···················· | ····················· | ···················· | ···················· | ···················· |
| K | 22 | hCoV-19/Australia/VIC11317/2020\|EPI_ISL_563327\|202 | ···················· | ···················· | ····················· | ···················· | ···················· | ···················· |
| K | 23 | hCoV-19/Australia/VIC7788/2020\|EPI_ISL_564658\|2020 | ···················· | ···················· | ····················· | ···················· | ···················· | ···················· |
| K | 24 | hCoV-19/Australia/VIC7790/2020\|EPI_ISL_564659\|2020 | ···················· | ···················· | ····················· | ···················· | ···················· | ···················· |
| K | 25 | hCoV-19/USA/MI-MDHHS-SC21929/2020\|EPI_ISL_565989\|2 | ···················· | ···················· | ····················· | ···················· | ···················· | ···················· |
| K | 26 | hCoV-19/England/ALDP-9BE8A6/2020\|EPI_ISL_568100\|20 | ···················· | ···················· | ····················· | ···················· | ···················· | ···················· |
| K | 27 | hCoV-19/England/ALDP-9BE026/2020\|EPI_ISL_568101\|20 | ···················· | ···················· | ····················· | ···················· | ···················· | ···················· |
| K | 28 | hCoV-19/England/ALDP-9BDA83/2020\|EPI_ISL_568102\|20 | ···················· | ···················· | ····················· | ···················· | ···················· | ···················· |
| K | 29 | hCoV-19/Wales/ALDP-9BE466/2020\|EPI_ISL_568103\|2020 | ···················· | ···················· | ····················· | ···················· | ···················· | ···················· |

K 30 hCoV-19/England/ALDP-9BC7C9/2020|EPI_ISL_568105|20 ···················· ·A·················· ····················· ···················· ···················· ···A················
K 31 hCoV-19/England/ALDP-9BE1AB/2020|EPI_ISL_568106|20 ···················· ···················· ····················· ···················· ···················· ····················
K 32 hCoV-19/England/ALDP-9BE76D/2020|EPI_ISL_568107|20 ···················· ···················· ····················· ···················· ···················· ····················
K 33 hCoV-19/England/ALDP-9BED4D/2020|EPI_ISL_568108|20 ···················· ···················· ····················· ···················· ···················· ····················
K 34 hCoV-19/England/ALDP-9BE9EF/2020|EPI_ISL_568109|20 ···················· ···················· ····················· ···················· ···················· ····················
K 35 hCoV-19/England/ALDP-9BDD6C/2020|EPI_ISL_568110|20 ···················· ···················· ····················· ···················· ···················· ····················
K 36 hCoV-19/England/ALDP-9BE730/2020|EPI_ISL_568111|20 ···················· ···················· ····················· ···················· ···················· ····················
K 37 hCoV-19/England/ALDP-9BDCC9/2020|EPI_ISL_568112|20 ···················· ···················· ····················· ···················· ···················· ····················
K 38 hCoV-19/England/ALDP-9BE93A/2020|EPI_ISL_568113|20 ···················· ···················· ····················· ···················· ···················· ····················
K 39 hCoV-19/England/ALDP-9BEADD/2020|EPI_ISL_568114|20 ···················· ···················· ····················· ···················· ···················· ····················
K 40 hCoV-19/England/ALDP-9BE7B8/2020|EPI_ISL_568115|20 ···················· ···················· ····················· ···················· ······T············· ····················
K 41 hCoV-19/Wales/ALDP-9BE1C9/2020|EPI_ISL_568116|2020 ···················· ···················· ····················· ···················· ···················· ····················
K 42 hCoV-19/England/ALDP-9BDDF3/2020|EPI_ISL_568118|20 ···················· ···················· ····················· ···················· ···················· ····················
K 43 hCoV-19/England/ALDP-9BC802/2020|EPI_ISL_568119|20 ···················· ···················· ····················· ···················· ···················· ····················
K 44 hCoV-19/England/ALDP-9BEB34/2020|EPI_ISL_568120|20 ···················· ···················· ····················· ···················· ···················· ····················
K 45 hCoV-19/England/ALDP-9BE202/2020|EPI_ISL_568121|20 ···················· ·A·················· ····················· ···················· ···················· ···A················
K 46 hCoV-19/England/ALDP-9BDBBD/2020|EPI_ISL_568123|20 ···················· ···················· ····················· ···················· ···················· ····················
K 47 hCoV-19/England/ALDP-9BECB9/2020|EPI_ISL_568124|20 ···················· ···················· ····················· ···················· ···················· ····················
K 48 hCoV-19/England/ALDP-9BE0CC/2020|EPI_ISL_568125|20 ···················· ····A··············· ····················· ···················· ···················· ····················
K 49 hCoV-19/England/ALDP-9BDD5D/2020|EPI_ISL_568126|20 ···················· ···················· ····················· ···················· ···················· ····················
K 50 hCoV-19/England/ALDP-9BE5BE/2020|EPI_ISL_568127|20 ···················· ···················· ····················· ···················· ···················· ····················
K 51 hCoV-19/England/ALDP-9BE1BA/2020|EPI_ISL_568128|20 ···················· ···················· ····················· ···················· ···················· ····················
K 52 hCoV-19/England/ALDP-9BDE2D/2020|EPI_ISL_568129|20 ···················· ···················· ····················· ···················· ···················· ····················
K 53 hCoV-19/England/ALDP-9BC8F3/2020|EPI_ISL_568130|20 ···················· ···················· ····················· ···················· ···················· ····················
K 54 hCoV-19/England/ALDP-9BDB44/2020|EPI_ISL_568131|20 ···················· ···················· ····················· ···················· ···················· ····················
K 55 hCoV-19/England/ALDP-9BDE1E/2020|EPI_ISL_568132|20 ···················· ···················· ····················· ···················· ···················· ····················
K 56 hCoV-19/England/ALDP-9BE90D/2020|EPI_ISL_568133|20 ···················· ···················· ····················· ···················· ···················· ····················
K 57 hCoV-19/England/ALDP-9BC8E4/2020|EPI_ISL_568134|20 ···················· ···················· ····················· ···················· ···················· ····················
K 58 hCoV-19/England/ALDP-9BDD11/2020|EPI_ISL_568135|20 ···················· ···················· ····················· ···················· ···················· ····················
K 59 hCoV-19/England/ALDP-9BEC9B/2020|EPI_ISL_568136|20 ···················· ···················· ····················· ···················· ···················· ····················
K 60 hCoV-19/England/ALDP-9BC7BA/2020|EPI_ISL_568137|20 ···················· ·A·················· ····················· ···················· ···················· ···A················
K 61 hCoV-19/England/ALDP-9BE7A9/2020|EPI_ISL_568138|20 ···················· ···················· ····················· ···················· ···················· ····················
K 62 hCoV-19/England/ALDP-9BE545/2020|EPI_ISL_568139|20 ···················· ···················· ····················· ···················· ···················· ····················
K 63 hCoV-19/England/ALDP-9BDB08/2020|EPI_ISL_568140|20 ···················· ···················· ····················· ···················· ···················· ····················
K 64 hCoV-19/England/ALDP-9BC86C/2020|EPI_ISL_568141|20 ···················· ···················· ····················· ···················· ···················· ····················
K 65 hCoV-19/England/ALDP-9BDF0C/2020|EPI_ISL_568142|20 ···················· ···················· ····················· ···················· ···················· ····················
K 66 hCoV-19/England/ALDP-9BE3E1/2020|EPI_ISL_568143|20 ···················· ···················· ····················· ···················· ···················· ····················
K 67 hCoV-19/England/ALDP-9BE9FE/2020|EPI_ISL_568144|20 ···················· ···················· ····················· ···················· ···················· ····················
K 68 hCoV-19/England/ALDP-9BE23F/2020|EPI_ISL_568146|20 ···················· ···················· ····················· ···················· ···················· ····················
K 69 hCoV-19/England/ALDP-9BDA74/2020|EPI_ISL_568147|20 ···················· ···················· ····················· ···················· ···················· ····················
K 70 hCoV-19/England/ALDP-9BC671/2020|EPI_ISL_568149|20 ···················· ···················· ····················· ···················· ···················· ····················
K 71 hCoV-19/England/ALDP-9BE220/2020|EPI_ISL_568150|20 ···················· ···················· ····················· ···················· ···················· ····················
K 72 hCoV-19/England/ALDP-9BE35A/2020|EPI_ISL_568151|20 ···················· ···················· ····················· ···················· ···················· ····················
K 73 hCoV-19/England/ALDP-9BE0EA/2020|EPI_ISL_568152|20 ···················· ····A··············· ····················· ···················· ···················· ····················
K 74 hCoV-19/England/ALDP-9BDB62/2020|EPI_ISL_568153|20 ···················· ···················· ····················· ···················· ···················· ····················
K 75 hCoV-19/England/ALDP-9BC8D5/2020|EPI_ISL_568154|20 ···················· ···················· ····················· ···················· ···················· ····················
K 76 hCoV-19/England/ALDP-9BE378/2020|EPI_ISL_568155|20 ···················· ···················· ····················· ···················· ···················· ····················
K 77 hCoV-19/England/ALDP-9BE2D5/2020|EPI_ISL_568156|20 ···················· ···················· ····················· ···················· ···················· ····················
K 78 hCoV-19/England/ALDP-9BC6CC/2020|EPI_ISL_568157|20 ···················· ···················· ····················· ···················· ···················· ····················
K 79 hCoV-19/England/ALDP-9BE0F9/2020|EPI_ISL_568158|20 ···················· ···················· ····················· ···················· ···················· ····················
K 80 hCoV-19/England/ALDP-9BC78D/2020|EPI_ISL_568159|20 ···················· ···················· ····················· ···················· ···················· ····················
K 81 hCoV-19/England/ALDP-9BC8B7/2020|EPI_ISL_568160|20 ···················· ···················· ····················· ···················· ···················· ····················
K 82 hCoV-19/England/ALDP-9BE6BB/2020|EPI_ISL_568161|20 ···················· ···················· ····················· ···················· ···················· ····················
K 83 hCoV-19/England/ALDP-9BDAED/2020|EPI_ISL_568162|20 ···················· ·A·················· ····················· ···················· ···················· ···A················
K 84 hCoV-19/England/ALDP-9BE4A2/2020|EPI_ISL_568163|20 ···················· ···················· ····················· ···················· ···················· ····················
K 85 hCoV-19/England/ALDP-9BEA19/2020|EPI_ISL_568164|20 ···················· ···················· ····················· ···················· ···················· ····················
K 86 hCoV-19/England/ALDP-9BE976/2020|EPI_ISL_568165|20 ···················· ···················· ····················· ···················· ···················· ····················
K 87 hCoV-19/England/ALDP-9BE17E/2020|EPI_ISL_568166|20 ···················· ···················· ····················· ···················· ···················· ····················
K 88 hCoV-19/England/ALDP-9BDA0B/2020|EPI_ISL_568167|20 ···················· ···················· ····················· ···················· ···················· ····················
K 89 hCoV-19/England/ALDP-9BD9E0/2020|EPI_ISL_568168|20 ···················· ···················· ····················· ···················· ···················· ····················
K 90 hCoV-19/England/ALDP-9BC978/2020|EPI_ISL_568169|20 ···················· ···················· ····················· ···················· ···················· ····················
K 91 hCoV-19/France/PAC-IHU-0237/2020|EPI_ISL_569400|20 ···················· ···················· ····················· ···················· ···················· ····················
K 92 hCoV-19/France/PAC-IHU-0238/2020|EPI_ISL_569401|20 ···················· ···················· ····················· ···················· ···················· ····················
K 93 hCoV-19/France/PAC-IHU-0239/2020|EPI_ISL_569402|20 ···················· ···················· ····················· ···················· ···················· ····················
K 94 hCoV-19/France/PAC-IHU-0241/2020|EPI_ISL_569403|20 ···················· ···················· ····················· ···················· ···················· ····················
K 95 hCoV-19/France/PAC-IHU-0242/2020|EPI_ISL_569404|20 ···················· ···················· ····················· ···················· ···················· ····················
K 96 hCoV-19/France/PAC-IHU-0243/2020|EPI_ISL_569405|20 ···················· ···················· ····················· ···················· ···················· ····················
K 97 hCoV-19/France/PAC-IHU-0246/2020|EPI_ISL_569406|20 ···················· ···················· ····················· ···················· ···················· ····················
K 98 hCoV-19/France/PAC-IHU-0247/2020|EPI_ISL_569407|20 ···················· ···················· ····················· ···················· ···················· ····················
K 99 hCoV-19/France/PAC-IHU-0248/2020|EPI_ISL_569408|20 ···················· ···················· ····················· ···················· ···················· ····················
K 100 hCoV-19/France/PAC-IHU-0250/2020|EPI_ISL_569410|20 ···················· ···················· ····················· ···················· ···················· ····················
K 101 hCoV-19/France/PAC-IHU-0251/2020|EPI_ISL_569411|20 ···················· ···················· ····················· ···················· ···················· ····················
K 102 hCoV-19/France/PAC-IHU-0252/2020|EPI_ISL_569412|20 ···················· ···················· ····················· ···················· ···················· ····················
K 103 hCoV-19/France/PAC-IHU-0254/2020|EPI_ISL_569413|20 ···················· ···················· ····················· ···················· ···················· ····················
K 104 hCoV-19/France/PAC-IHU-0256/2020|EPI_ISL_569414|20 ···················· ···················· ····················· ···················· ···················· ····················
K 105 hCoV-19/France/PAC-IHU-0257/2020|EPI_ISL_569415|20 ···················· ···················· ····················· ···················· ···················· ····················
K 106 hCoV-19/France/PAC-IHU-0258/2020|EPI_ISL_569416|20 ···················· ···················· ····················· ···················· ···················· ····················
K 107 hCoV-19/France/PAC-IHU-0259/2020|EPI_ISL_569417|20 ···················· ···················· ····················· ···················· ···················· ····················
K 108 hCoV-19/France/PAC-IHU-0261/2020|EPI_ISL_569418|20 ···················· ···················· ····················· ···················· ······T············· ····················
K 109 hCoV-19/France/PAC-IHU-0262/2020|EPI_ISL_569419|20 ···················· ···················· ····················· ···················· ···················· ····················

K 110 hCoV-19/France/PAC-IHU-0263/2020|EPI_ISL_569420|20 ···················· ···················· ···················· ···················· ···················· ····················
K 111 hCoV-19/France/PAC-IHU-0266/2020|EPI_ISL_569421|20 ···················· ···················· ···················· ···················· ···················· ····················
K 112 hCoV-19/France/PAC-IHU-0268/2020|EPI_ISL_569422|20 ···················· ···················· ···················· ···················· ···················· ····················
K 113 hCoV-19/France/PAC-IHU-0269/2020|EPI_ISL_569423|20 ···················· ···················· ···················· ···················· ···················· ····················
K 114 hCoV-19/France/PAC-IHU-0271/2020|EPI_ISL_569424|20 ···················· ···················· ···················· ···················· ···················· ····················
K 115 hCoV-19/France/PAC-IHU-0274/2020|EPI_ISL_569425|20 ···················· ···················· ···················· ···················· ···················· ····················
K 116 hCoV-19/France/PAC-IHU-0276/2020|EPI_ISL_569426|20 ···················· ···················· ···················· ···················· ···················· ····················
K 117 hCoV-19/France/PAC-IHU-0280/2020|EPI_ISL_569427|20 ···················· ···················· ···················· ···················· ···················· ····················
K 118 hCoV-19/France/PAC-IHU-0281/2020|EPI_ISL_569428|20 ···················· ···················· ···················· ···················· ···················· ····················
K 119 hCoV-19/France/PAC-IHU-0282/2020|EPI_ISL_569429|20 ···················· ···················· ···················· ···················· ···················· ····················
K 120 hCoV-19/France/PAC-IHU-0284/2020|EPI_ISL_569431|20 ···················· ···················· ···················· ···················· ···················· ····················
K 121 hCoV-19/France/PAC-IHU-0286/2020|EPI_ISL_569432|20 ···················· ···················· ···················· ···················· ···················· ····················
K 122 hCoV-19/France/PAC-IHU-0287/2020|EPI_ISL_569433|20 ···················· ···················· ···················· ···················· ···················· ····················
K 123 hCoV-19/France/PAC-IHU-0288/2020|EPI_ISL_569434|20 ···················· ···················· ···················· ···················· ···················· ····················
K 124 hCoV-19/France/PAC-IHU-0290/2020|EPI_ISL_569435|20 ···················· ···················· ···················· ···················· ···················· ····················
K 125 hCoV-19/France/PAC-IHU-0291/2020|EPI_ISL_569436|20 ···················· ···················· ···················· ···················· ···················· ····················
K 126 hCoV-19/France/PAC-IHU-0293/2020|EPI_ISL_569437|20 ···················· ···················· ···················· ···················· ···················· ····················
K 127 hCoV-19/France/PAC-IHU-0295/2020|EPI_ISL_569438|20 ···················· ···················· ···················· ···················· ···················· ····················
K 128 hCoV-19/France/PAC-IHU-0297/2020|EPI_ISL_569440|20 ···················· ···················· ···················· ···················· ···················· ····················
K 129 hCoV-19/France/PAC-IHU-0298/2020|EPI_ISL_569441|20 ···················· ···················· ···················· ···················· ···················· ····················
K 130 hCoV-19/France/PAC-IHU-0300/2020|EPI_ISL_569443|20 ···················· ···················· ···················· ···················· ···················· ····················
K 131 hCoV-19/France/PAC-IHU-0303/2020|EPI_ISL_569444|20 ···················· ···················· ···················· ···················· ···················· ····················
K 132 hCoV-19/France/PAC-IHU-0304/2020|EPI_ISL_569445|20 ···················· ···················· ···················· ···················· ···················· ····················
K 133 hCoV-19/France/PAC-IHU-0306/2020|EPI_ISL_569446|20 ···················· ···················· ···················· ···················· ···················· ····················
K 134 hCoV-19/France/PAC-IHU-0308/2020|EPI_ISL_569447|20 ···················· ···················· ···················· ···················· ···················· ····················
K 135 hCoV-19/France/PAC-IHU-0309/2020|EPI_ISL_569448|20 ···················· ···················· ···················· ···················· ···················· ····················
K 136 hCoV-19/France/PAC-IHU-0316/2020|EPI_ISL_569449|20 ···················· ···················· ···················· ···················· ···················· ····················
K 137 hCoV-19/France/PAC-IHU-0318/2020|EPI_ISL_569450|20 ···················· ···················· ···················· ···················· ···················· ····················
K 138 hCoV-19/France/PAC-IHU-0319/2020|EPI_ISL_569451|20 ···················· ···················· ···················· ···················· ···················· ····················
K 139 hCoV-19/France/PAC-IHU-0327/2020|EPI_ISL_569456|20 ···················· ···················· ···················· ···················· ···················· ····················
K 140 hCoV-19/France/PAC-IHU-0328/2020|EPI_ISL_569457|20 ···················· ···················· ···················· ···················· ···················· ····················
K 141 hCoV-19/France/PAC-IHU-0329/2020|EPI_ISL_569458|20 ···················· ···················· ···················· ···················· ···················· ····················
K 142 hCoV-19/France/PAC-IHU-0332/2020|EPI_ISL_569459|20 ···················· ····A··············· ···················· ···················· ···················· ····················
K 143 hCoV-19/France/PAC-IHU-0333/2020|EPI_ISL_569460|20 ···················· ···················· ···················· ···················· ···················· ····················
K 145 hCoV-19/France/PAC-IHU-0336/2020|EPI_ISL_569462|20 ···················· ···················· ···················· ···················· ···················· ····················
K 146 hCoV-19/France/PAC-IHU-0337/2020|EPI_ISL_569463|20 ···················· ···················· ···················· ···················· ···················· ····················
K 147 hCoV-19/France/PAC-IHU-0338/2020|EPI_ISL_569464|20 ···················· ···················· ···················· ···················· ···················· ····················
K 148 hCoV-19/France/PAC-IHU-0340/2020|EPI_ISL_569465|20 ···················· ···················· ···················· ···················· ···················· ····················
K 149 hCoV-19/France/PAC-IHU-0342/2020|EPI_ISL_569466|20 ···················· ···················· ···················· ···················· ···················· ····················
K 150 hCoV-19/France/PAC-IHU-0344/2020|EPI_ISL_569467|20 ···················· ···················· ···················· ···················· ···················· ····················
K 151 hCoV-19/France/PAC-IHU-0345/2020|EPI_ISL_569468|20 ···················· ···················· ···················· ···················· ···················· ····················
K 152 hCoV-19/France/PAC-IHU-0347/2020|EPI_ISL_569469|20 ···················· ···················· ···················· ···················· ···················· ····················
K 153 hCoV-19/France/PAC-IHU-0349/2020|EPI_ISL_569470|20 ···················· ···················· ···················· ···················· ···················· ····················
K 154 hCoV-19/France/PAC-IHU-0351/2020|EPI_ISL_569471|20 ···················· ··········A········· ···················· ···················· ···················· ····················
K 155 hCoV-19/France/PAC-IHU-0354/2020|EPI_ISL_569472|20 ···················· ···················· ···················· ···················· ···················· ····················
K 156 hCoV-19/France/PAC-IHU-0355/2020|EPI_ISL_569473|20 ···················· ···················· ···················· ···················· ···················· ····················
K 157 hCoV-19/France/PAC-IHU-0361/2020|EPI_ISL_569474|20 ···················· ···················· ···················· ···················· ···················· ····················
K 158 hCoV-19/France/PAC-IHU-0366/2020|EPI_ISL_569475|20 ···················· ···················· ···················· ···················· ···················· ····················
K 159 hCoV-19/France/PAC-IHU-0369/2020|EPI_ISL_569476|20 ···················· ···················· ···················· ···················· ···················· ····················
K 160 hCoV-19/France/PAC-IHU-0375/2020|EPI_ISL_569477|20 ···················· ···················· ···················· ···················· ···················· ····················
K 161 hCoV-19/France/PAC-IHU-0379/2020|EPI_ISL_569478|20 ···················· ···················· ···················· ···················· ···················· ····················
K 162 hCoV-19/France/PAC-IHU-0381/2020|EPI_ISL_569479|20 ···················· ···················· ···················· ···················· ···················· ····················
K 163 hCoV-19/France/PAC-IHU-0385/2020|EPI_ISL_569481|20 ···················· ···················· ···················· ···················· ···················· ····················
K 164 hCoV-19/France/PAC-IHU-0388/2020|EPI_ISL_569483|20 ···················· ···················· ···················· ···················· ···················· ····················
K 165 hCoV-19/France/PAC-IHU-0389/2020|EPI_ISL_569484|20 ···················· ···················· ···················· ···················· ···················· ····················
K 166 hCoV-19/France/PAC-IHU-0391/2020|EPI_ISL_569485|20 ···················· ···················· ···················· ···················· ···················· ····················
K 167 hCoV-19/France/PAC-IHU-0400/2020|EPI_ISL_569489|20 ···················· ···················· ···················· ···················· ···················· ····················
K 168 hCoV-19/France/PAC-IHU-0401/2020|EPI_ISL_569490|20 ···················· ···················· ···················· ···················· ···················· ····················
K 169 hCoV-19/France/PAC-IHU-0405/2020|EPI_ISL_569491|20 ···················· ···················· ···················· ···················· ···················· ····················
K 170 hCoV-19/France/PAC-IHU-0407/2020|EPI_ISL_569492|20 ···················· ···················· ···················· ···················· ···················· ····················
K 171 hCoV-19/France/PAC-IHU-0409/2020|EPI_ISL_569493|20 ···················· ···················· ···················· ···················· ···················· ····················
K 172 hCoV-19/France/PAC-IHU-0411/2020|EPI_ISL_569494|20 ···················· ····T··············· ···················· ···················· ···················· ····················
K 173 hCoV-19/France/PAC-IHU-0412/2020|EPI_ISL_569495|20 ···················· ···················· ···················· ···················· ···················· ····················
K 174 hCoV-19/France/PAC-IHU-0417/2020|EPI_ISL_569496|20 ···················· ···················· ···················· ···················· ···················· ····················
K 175 hCoV-19/France/PAC-IHU-0419/2020|EPI_ISL_569497|20 ···················· ···················· ···················· ···················· ···················· ····················
K 176 hCoV-19/France/PAC-IHU-0423/2020|EPI_ISL_569498|20 ···················· ···················· ···················· ···················· ···················· ····················
K 177 hCoV-19/France/PAC-IHU-0424/2020|EPI_ISL_569499|20 ···················· ···················· ···················· ···················· ···················· ····················
K 178 hCoV-19/USA/WA-UW-11350/2020|EPI_ISL_570400|2020-0 ···················· ···················· ···················· ···················· ···················· ····················
K 179 hCoV-19/USA/WA-UW-11391/2020|EPI_ISL_570401|2020-0 ···················· ···················· ···················· ···················· ···················· ····················
K 180 hCoV-19/USA/WA-UW-11394/2020|EPI_ISL_570402|2020-0 ···················· ···················· ···················· ···················· ···················· ····················
K 181 hCoV-19/USA/WA-UW-11401/2020|EPI_ISL_570403|2020-0 ···················· ···················· ···················· ···················· ···················· ····················
K 182 hCoV-19/USA/WA-UW-11425/2020|EPI_ISL_570404|2020-0 ···················· ·A·················· ···················· ···················· ···················· ····················
K 183 hCoV-19/USA/WA-UW-11436/2020|EPI_ISL_570405|2020-0 ···················· ···················· ···················· ···················· ···················· ····················
K 184 hCoV-19/USA/WA-UW-11437/2020|EPI_ISL_570406|2020-0 ···················· ···················· ···················· ···················· ···················· ····················
K 185 hCoV-19/USA/WA-UW-11438/2020|EPI_ISL_570407|2020-0 ···················· ···················· ···················· ···················· ···················· ····················
K 186 hCoV-19/USA/WA-UW-11442/2020|EPI_ISL_570408|2020-0 ···················· ···················· ···················· ···················· ···················· ····················
K 187 hCoV-19/USA/WA-UW-11443/2020|EPI_ISL_570409|2020-0 ···················· ·A·················· ···················· ···················· ···················· ····················
K 188 hCoV-19/USA/WA-UW-11447/2020|EPI_ISL_570410|2020-0 ···················· ···················· ···················· ···················· ···················· ····················
K 189 hCoV-19/USA/WA-UW-11449/2020|EPI_ISL_570411|2020-0 ···················· ···················· ···················· ···················· ···················· ····················
K 190 hCoV-19/USA/WA-UW-11453/2020|EPI_ISL_570412|2020-0 ···················· ···················· ···················· ···················· ···················· ····················

| | | | | | | | | |
|---|---|---|---|---|---|---|---|---|
| K | 191 | hCoV-19/USA/WA-UW-11454/2020\|EPI_ISL_570413\|2020-0 | ···················· | ···················· | ····················· | ···················· | ···················· | ···················· |
| K | 192 | hCoV-19/USA/WA-UW-11459/2020\|EPI_ISL_570414\|2020-0 | ···················· | ···················· | ····················· | ···················· | ···················· | ···················· |
| K | 193 | hCoV-19/USA/WA-UW-11472/2020\|EPI_ISL_570415\|2020-0 | ···················· | ···················· | ····················· | ···················· | ···················· | ···················· |
| K | 194 | hCoV-19/USA/WA-UW-11478/2020\|EPI_ISL_570416\|2020-0 | ···················· | ·A·················· | ····················· | ···················· | ···················· | ···················· |
| K | 195 | hCoV-19/USA/WA-UW-11480/2020\|EPI_ISL_570417\|2020-0 | ···················· | ···················· | ····················· | ···················· | ···················· | ···················· |
| K | 196 | hCoV-19/USA/WA-UW-11481/2020\|EPI_ISL_570418\|2020-0 | ···················· | ···················· | ····················· | ···················· | ···················· | ···················· |
| K | 197 | hCoV-19/USA/WA-UW-11484/2020\|EPI_ISL_570419\|2020-0 | ···················· | ·A·················· | ····················· | ···················· | ···················· | ···················· |
| K | 198 | hCoV-19/USA/WA-UW-11487/2020\|EPI_ISL_570420\|2020-0 | ···················· | ···················· | ····················· | ···················· | ···················· | ···················· |
| K | 199 | hCoV-19/USA/WA-UW-11488/2020\|EPI_ISL_570421\|2020-0 | ···················· | ···················· | ····················· | ···················· | ···················· | ···················· |
| K | 200 | hCoV-19/USA/WA-UW-11489/2020\|EPI_ISL_570422\|2020-0 | ···················· | ···················· | ····················· | ···················· | ···················· | ···················· |
| K | 201 | hCoV-19/USA/WA-UW-11499/2020\|EPI_ISL_570423\|2020-0 | ···················· | ·A·················· | ····················· | ···················· | ···················· | ···················· |
| K | 202 | hCoV-19/USA/WA-UW-11512/2020\|EPI_ISL_570424\|2020-0 | ···················· | ·A·················· | ····················· | ···················· | ···················· | ···················· |
| K | 203 | hCoV-19/USA/WA-UW-11518/2020\|EPI_ISL_570426\|2020-0 | ···················· | ···················· | ····················· | ···················· | ···················· | ···················· |
| K | 204 | hCoV-19/USA/WA-UW-11519/2020\|EPI_ISL_570427\|2020-0 | ···················· | ···················· | ····················· | ···················· | ···················· | ···················· |
| K | 205 | hCoV-19/USA/WA-UW-11521/2020\|EPI_ISL_570428\|2020-0 | ···················· | ···················· | ····················· | ···················· | ···················· | ···················· |
| K | 206 | hCoV-19/USA/WA-UW-11526/2020\|EPI_ISL_570429\|2020-0 | ···················· | ·A·················· | ····················· | ···················· | ···················· | ···················· |
| K | 207 | hCoV-19/USA/WA-UW-11529/2020\|EPI_ISL_570430\|2020-0 | ···················· | ···················· | ····················· | ···················· | ···················· | ···················· |
| K | 208 | hCoV-19/USA/WA-UW-11530/2020\|EPI_ISL_570431\|2020-0 | ···················· | ···················· | ····················· | ···················· | ···················· | ···················· |
| K | 209 | hCoV-19/USA/WA-UW-11533/2020\|EPI_ISL_570432\|2020-0 | ···················· | ···················· | ····················· | ···················· | ···················· | ···················· |
| K | 210 | hCoV-19/USA/WA-UW-11537/2020\|EPI_ISL_570433\|2020-0 | ···················· | ···················· | ····················· | ···················· | ···················· | ···················· |
| K | 211 | hCoV-19/USA/WA-UW-11544/2020\|EPI_ISL_570434\|2020-0 | ···················· | ···················· | ····················· | ···················· | ···················· | ···················· |
| K | 212 | hCoV-19/USA/WA-UW-11556/2020\|EPI_ISL_570435\|2020-0 | ···················· | ···················· | ····················· | ···················· | ···················· | ···················· |
| K | 213 | hCoV-19/USA/WA-UW-11561/2020\|EPI_ISL_570436\|2020-0 | ···················· | ···················· | ····················· | ···················· | ···················· | ···················· |
| K | 214 | hCoV-19/USA/WA-UW-11562/2020\|EPI_ISL_570437\|2020-0 | ···················· | ···················· | ····················· | ···················· | ···················· | ···················· |
| K | 215 | hCoV-19/USA/WA-UW-11564/2020\|EPI_ISL_570438\|2020-0 | ···················· | ···················· | ····················· | ···················· | ···················· | ···················· |
| K | 216 | hCoV-19/USA/WA-UW-11571/2020\|EPI_ISL_570439\|2020-0 | ···················· | ···················· | ····················· | ···················· | ···················· | ···················· |
| K | 217 | hCoV-19/USA/WA-UW-11572/2020\|EPI_ISL_570440\|2020-0 | ···················· | ···················· | ····················· | ···················· | ···················· | ···················· |
| K | 218 | hCoV-19/USA/WA-UW-11576/2020\|EPI_ISL_570441\|2020-0 | ···················· | ···················· | ····················· | ···················· | ···················· | ···················· |
| K | 219 | hCoV-19/USA/WA-UW-11580/2020\|EPI_ISL_570442\|2020-0 | ···················· | ···················· | ····················· | ···················· | ···················· | ···················· |
| K | 220 | hCoV-19/USA/WA-UW-11592/2020\|EPI_ISL_570444\|2020-0 | ···················· | ···················· | ····················· | ···················· | ···················· | ···················· |
| K | 221 | hCoV-19/USA/WA-UW-11602/2020\|EPI_ISL_570445\|2020-0 | ···················· | ···················· | ····················· | ···················· | ···················· | ···················· |
| K | 222 | hCoV-19/USA/WA-UW-11608/2020\|EPI_ISL_570446\|2020-0 | ···················· | ···················· | ····················· | ···················· | ···················· | ···················· |
| K | 223 | hCoV-19/USA/WA-UW-11611/2020\|EPI_ISL_570447\|2020-0 | ···················· | ···················· | ····················· | ···················· | ···················· | ···················· |
| K | 224 | hCoV-19/USA/WA-UW-11630/2020\|EPI_ISL_570449\|2020-0 | ···················· | ···················· | ····················· | ···················· | ···················· | ···················· |
| K | 225 | hCoV-19/USA/WA-UW-11633/2020\|EPI_ISL_570450\|2020-0 | ···················· | ···················· | ····················· | ···················· | ···················· | ···················· |
| K | 226 | hCoV-19/USA/WA-UW-11635/2020\|EPI_ISL_570451\|2020-0 | ···················· | ···················· | ····················· | ···················· | ···················· | ···················· |
| K | 227 | hCoV-19/USA/WA-UW-11640/2020\|EPI_ISL_570452\|2020-0 | ···················· | ···················· | ····················· | ···················· | ···················· | ···················· |
| K | 228 | hCoV-19/USA/WA-UW-11645/2020\|EPI_ISL_570453\|2020-0 | ···················· | ···················· | ····················· | ···················· | ···················· | ···················· |
| K | 229 | hCoV-19/USA/WA-UW-11648/2020\|EPI_ISL_570454\|2020-0 | ···················· | ···················· | ····················· | ···················· | ···················· | ···················· |
| K | 230 | hCoV-19/USA/WA-UW-11656/2020\|EPI_ISL_570455\|2020-0 | ···················· | ···················· | ····················· | ···················· | ···················· | ···················· |
| K | 231 | hCoV-19/USA/WA-UW-11661/2020\|EPI_ISL_570456\|2020-0 | ···················· | ···················· | ····················· | ···················· | ···················· | ···················· |
| K | 232 | hCoV-19/USA/WA-UW-11664/2020\|EPI_ISL_570457\|2020-0 | ···················· | ···················· | ····················· | ···················· | ······T············· | ···················· |
| K | 233 | hCoV-19/USA/WA-UW-11671/2020\|EPI_ISL_570458\|2020-0 | ···················· | ···················· | ····················· | ···················· | ···················· | ···················· |
| K | 234 | hCoV-19/USA/WA-UW-11673/2020\|EPI_ISL_570459\|2020-0 | ···················· | ···················· | ····················· | ···················· | ···················· | ···················· |
| K | 235 | hCoV-19/USA/WA-UW-11703/2020\|EPI_ISL_570460\|2020-0 | ···················· | ···················· | ····················· | ···················· | ···················· | ···················· |
| K | 236 | hCoV-19/USA/WA-UW-11709/2020\|EPI_ISL_570461\|2020-0 | ···················· | ···················· | ····················· | ···················· | ···················· | ···················· |
| K | 237 | hCoV-19/USA/WA-UW-11710/2020\|EPI_ISL_570462\|2020-0 | ···················· | ·A·················· | ····················· | ···················· | ···················· | ···················· |
| K | 238 | hCoV-19/USA/WA-UW-11722/2020\|EPI_ISL_570463\|2020-0 | ···················· | ···················· | ····················· | ···················· | ···················· | ···················· |
| K | 239 | hCoV-19/USA/WA-UW-11728/2020\|EPI_ISL_570464\|2020-0 | ···················· | ···················· | ····················· | ···················· | ···················· | ···················· |
| K | 240 | hCoV-19/USA/OR-UW-11742/2020\|EPI_ISL_570465\|2020-0 | ···················· | ·A·················· | ····················· | ···················· | ···················· | ···················· |
| K | 241 | hCoV-19/USA/WA-UW-11761/2020\|EPI_ISL_570466\|2020-0 | ···················· | ·A·················· | ····················· | ···················· | ···················· | ···················· |
| K | 242 | hCoV-19/USA/WA-UW-11762/2020\|EPI_ISL_570467\|2020-0 | ···················· | ···················· | ····················· | ···················· | ···················· | ···················· |
| K | 243 | hCoV-19/USA/WA-UW-11763/2020\|EPI_ISL_570468\|2020-0 | ···················· | ···················· | ····················· | ···················· | ···················· | ···················· |
| K | 244 | hCoV-19/USA/WA-UW-11764/2020\|EPI_ISL_570469\|2020-0 | ···················· | ···················· | ····················· | ···················· | ···················· | ···················· |
| K | 245 | hCoV-19/USA/WA-UW-11765/2020\|EPI_ISL_570470\|2020-0 | ···················· | ···················· | ····················· | ···················· | ···················· | ···················· |
| K | 246 | hCoV-19/USA/WA-UW-11775/2020\|EPI_ISL_570471\|2020-0 | ···················· | ···················· | ····················· | ···················· | ···················· | ···················· |
| K | 247 | hCoV-19/USA/WA-UW-11780/2020\|EPI_ISL_570472\|2020-0 | ···················· | ···················· | ····················· | ···················· | ···················· | ···················· |
| K | 248 | hCoV-19/USA/WA-UW-11813/2020\|EPI_ISL_570473\|2020-0 | ···················· | ···················· | ····················· | ···················· | ···················· | ···················· |
| K | 249 | hCoV-19/USA/WA-UW-11823/2020\|EPI_ISL_570474\|2020-0 | ···················· | ·A·················· | ····················· | ···················· | ···················· | ···················· |
| K | 250 | hCoV-19/USA/WA-UW-11836/2020\|EPI_ISL_570475\|2020-0 | ···················· | ···················· | ····················· | ···················· | ···················· | ···················· |
| K | 251 | hCoV-19/USA/WA-UW-11903/2020\|EPI_ISL_570476\|2020-0 | ···················· | ···················· | ····················· | ···················· | ···················· | ···················· |
| K | 252 | hCoV-19/USA/WA-UW-11909/2020\|EPI_ISL_570478\|2020-0 | ···················· | ···················· | ····················· | ···················· | ···················· | ···················· |
| K | 253 | hCoV-19/USA/WA-UW-11919/2020\|EPI_ISL_570480\|2020-0 | ···················· | ·A·················· | ····················· | ···················· | ···················· | ···················· |
| K | 254 | hCoV-19/USA/WA-UW-11989/2020\|EPI_ISL_570481\|2020-0 | ···················· | ···················· | ····················· | ···················· | ···················· | ···················· |
| K | 255 | hCoV-19/USA/WA-UW-12068/2020\|EPI_ISL_570482\|2020-0 | ···················· | ···················· | ····················· | ···················· | ···················· | ···················· |
| K | 256 | hCoV-19/USA/WA-UW-12069/2020\|EPI_ISL_570483\|2020-0 | ···················· | ·A·················· | ····················· | ···················· | ···················· | ···················· |
| K | 257 | hCoV-19/Australia/VIC11217/2020\|EPI_ISL_563301\|202 | ···················· | ···················· | ····················· | ···················· | ···················· | ···················· |
| K | 258 | hCoV-19/Australia/VIC11220/2020\|EPI_ISL_563302\|202 | ···················· | ···················· | ····················· | ···················· | ···················· | ···················· |
| K | 260 | hCoV-19/Australia/VIC11224/2020\|EPI_ISL_563304\|202 | ···················· | ···················· | ····················· | ···················· | ···················· | ···················· |
| K | 261 | hCoV-19/Australia/VIC11228/2020\|EPI_ISL_563305\|202 | ···················· | ···················· | ····················· | ···················· | ···················· | ···················· |
| K | 262 | hCoV-19/Australia/VIC11230/2020\|EPI_ISL_563306\|202 | ···················· | ···················· | ····················· | ···················· | ···················· | ···················· |
| K | 264 | hCoV-19/Australia/VIC11241/2020\|EPI_ISL_563308\|202 | ···················· | ···················· | ····················· | ···················· | ···················· | ···················· |
| K | 265 | hCoV-19/Australia/VIC11243/2020\|EPI_ISL_563309\|202 | ···················· | ···················· | ····················· | ···················· | ···················· | ···················· |
| K | 266 | hCoV-19/Australia/VIC11254/2020\|EPI_ISL_563310\|202 | ···················· | ···················· | ····················· | ···················· | ···················· | ···················· |
| K | 267 | hCoV-19/Australia/VIC11257/2020\|EPI_ISL_563311\|202 | ···················· | ···················· | ····················· | ···················· | ···················· | ···················· |
| K | 268 | hCoV-19/Australia/VIC11283/2020\|EPI_ISL_563312\|202 | ···················· | ···················· | ····················· | ···················· | ···················· | ···················· |
| K | 269 | hCoV-19/Australia/VIC11284/2020\|EPI_ISL_563313\|202 | ···················· | ···················· | ····················· | ···················· | ···················· | ···················· |
| K | 270 | hCoV-19/Australia/VIC11285/2020\|EPI_ISL_563314\|202 | ···················· | ···················· | ····················· | ···················· | ···················· | ···················· |
| K | 271 | hCoV-19/Australia/VIC11286/2020\|EPI_ISL_563315\|202 | ···················· | ···················· | ····················· | ···················· | ···················· | ···················· |
| K | 272 | hCoV-19/Australia/VIC11287/2020\|EPI_ISL_563316\|202 | ···················· | ···················· | ····················· | ···················· | ···················· | ···················· |

| | | | | | | | | | |
|---|---|---|---|---|---|---|---|---|---|
| K | 273 | hCoV-19/Australia/VIC11288/2020\|EPI_ISL_563317\|202 | ···················· | ···················· | ···················· | ···················· | ···················· | ···················· | |
| K | 274 | hCoV-19/Australia/VIC11290/2020\|EPI_ISL_563318\|202 | ···················· | ···················· | ···················· | ···················· | ···················· | ···················· | |
| K | 275 | hCoV-19/Australia/VIC11291/2020\|EPI_ISL_563319\|202 | ···················· | ···················· | ···················· | ···················· | ···················· | ···················· | |
| K | 276 | hCoV-19/Australia/VIC11294/2020\|EPI_ISL_563320\|202 | ···················· | ···················· | ···················· | ···················· | ···················· | ···················· | |
| K | 277 | hCoV-19/Australia/VIC11295/2020\|EPI_ISL_563321\|202 | ···················· | ···················· | ···················· | ···················· | ···················· | ···················· | |
| K | 279 | hCoV-19/Australia/VIC11304/2020\|EPI_ISL_563323\|202 | ···················· | ···················· | ···················· | ···················· | ···················· | ···················· | |
| K | 280 | hCoV-19/Australia/VIC11305/2020\|EPI_ISL_563324\|202 | ···················· | ···················· | ···················· | ···················· | ···················· | ···················· | |
| K | 281 | hCoV-19/Australia/VIC11312/2020\|EPI_ISL_563325\|202 | ···················· | ···················· | ···················· | ···················· | ···················· | ···················· | |
| K | 282 | hCoV-19/Australia/VIC11316/2020\|EPI_ISL_563326\|202 | ···················· | ···················· | ···················· | ···················· | ···················· | ···················· | |
| K | 283 | hCoV-19/Australia/VIC11320/2020\|EPI_ISL_563328\|202 | ···················· | ···················· | ···················· | ···················· | ···················· | ···················· | |
| K | 284 | hCoV-19/Australia/VIC11323/2020\|EPI_ISL_563329\|202 | ···················· | ···················· | ···················· | ···················· | ···················· | ···················· | |
| K | 285 | hCoV-19/Australia/VIC11324/2020\|EPI_ISL_563330\|202 | ···················· | ···················· | ···················· | ···················· | ···················· | ···················· | |
| K | 286 | hCoV-19/Australia/VIC11329/2020\|EPI_ISL_563331\|202 | ···················· | ···················· | ···················· | ···················· | ·················N·· | ···················· | 1 |
| K | 287 | hCoV-19/Australia/VIC11335/2020\|EPI_ISL_563332\|202 | ···················· | ···················· | ···················· | ···················· | ·················N·· | ···················· | 1 |
| K | 288 | hCoV-19/Australia/VIC11336/2020\|EPI_ISL_563333\|202 | ···················· | ···················· | ···················· | ···················· | ···················· | ···················· | |
| K | 290 | hCoV-19/Australia/VIC11338/2020\|EPI_ISL_563335\|202 | ···················· | ···················· | ···················· | ···················· | ···················· | ···················· | |
| K | 291 | hCoV-19/Australia/VIC11339/2020\|EPI_ISL_563336\|202 | ···················· | ···················· | ···················· | ···················· | ···················· | ···················· | |
| K | 292 | hCoV-19/Australia/VIC11340/2020\|EPI_ISL_563337\|202 | ···················· | ···················· | ···················· | ···················· | ···················· | ···················· | |
| K | 294 | hCoV-19/Australia/VIC11343/2020\|EPI_ISL_563339\|202 | ···················· | ···················· | ···················· | ···················· | ···················· | ···················· | |
| K | 295 | hCoV-19/Australia/VIC11344/2020\|EPI_ISL_563340\|202 | ···················· | ···················· | ···················· | ···················· | ···················· | ···················· | |
| K | 296 | hCoV-19/Australia/VIC11349/2020\|EPI_ISL_563341\|202 | ···················· | ···················· | ···················· | ···················· | ···················· | ···················· | |
| K | 297 | hCoV-19/Australia/VIC11352/2020\|EPI_ISL_563342\|202 | ···················· | ···················· | ···················· | ···················· | ···················· | ···················· | |
| K | 298 | hCoV-19/Australia/VIC11353/2020\|EPI_ISL_563343\|202 | ···················· | ···················· | ···················· | ···················· | ···················· | ···················· | |
| K | 299 | hCoV-19/Australia/VIC11357/2020\|EPI_ISL_563344\|202 | ···················· | ···················· | ···················· | ···················· | ···················· | ···················· | |
| K | 300 | hCoV-19/Australia/VIC11364/2020\|EPI_ISL_563345\|202 | ···················· | ···················· | ···················· | ···················· | ···················· | ···················· | |
| K | 301 | hCoV-19/Australia/VIC11369/2020\|EPI_ISL_563346\|202 | ···················· | ···················· | ···················· | ···················· | ·N·················· | ···················· | |
| K | 303 | hCoV-19/Australia/VIC11378/2020\|EPI_ISL_563348\|202 | ···················· | ···················· | ···················· | ···················· | ···················· | ···················· | |
| K | 304 | hCoV-19/Australia/VIC11379/2020\|EPI_ISL_563349\|202 | ···················· | ···················· | ···················· | ···················· | ···················· | ···················· | |
| K | 305 | hCoV-19/Australia/VIC11387/2020\|EPI_ISL_563350\|202 | ···················· | ···················· | ···················· | ···················· | ···················· | ···················· | |
| K | 306 | hCoV-19/Australia/VIC11388/2020\|EPI_ISL_563351\|202 | ···················· | ···················· | ···················· | ···················· | ···················· | ···················· | |
| K | 307 | hCoV-19/Australia/VIC11391/2020\|EPI_ISL_563352\|202 | ···················· | ···················· | ···················· | ···················· | ···················· | ···················· | |
| K | 308 | hCoV-19/Australia/VIC11392/2020\|EPI_ISL_563353\|202 | ···················· | ···················· | ···················· | ···················· | ···················· | ···················· | |
| K | 310 | hCoV-19/Australia/VIC11396/2020\|EPI_ISL_563355\|202 | ···················· | ···················· | ···················· | ···················· | ···················· | ···················· | |
| K | 312 | hCoV-19/Australia/VIC11399/2020\|EPI_ISL_563357\|202 | ···················· | ···················· | ···················· | ···················· | ···················· | ···················· | |
| K | 313 | hCoV-19/Australia/VIC11402/2020\|EPI_ISL_563358\|202 | ···················· | ···················· | ···················· | ···················· | ···················· | ···················· | |
| K | 314 | hCoV-19/Australia/VIC11403/2020\|EPI_ISL_563359\|202 | ···················· | ···················· | ···················· | ···················· | ···················· | ···················· | |
| K | 315 | hCoV-19/Australia/VIC11410/2020\|EPI_ISL_563360\|202 | ···················· | ···················· | ···················· | ···················· | ···················· | ·················M·· | 1 |
| K | 317 | hCoV-19/Australia/VIC11413/2020\|EPI_ISL_563362\|202 | ···················· | ···················· | ···················· | ···················· | ···················· | ···················· | |
| K | 318 | hCoV-19/Australia/VIC11414/2020\|EPI_ISL_563363\|202 | ···················· | ···················· | ···················· | ···················· | ···················· | ···················· | |
| K | 319 | hCoV-19/Australia/VIC11415/2020\|EPI_ISL_563364\|202 | ···················· | ···················· | ···················· | ···················· | ···················· | ···················· | |
| K | 320 | hCoV-19/Australia/VIC11418/2020\|EPI_ISL_563365\|202 | ···················· | ···················· | ···················· | ···················· | ···················· | ···················· | |
| K | 321 | hCoV-19/Australia/VIC11420/2020\|EPI_ISL_563366\|202 | ···················· | ···················· | ···················· | ···················· | ···················· | ···················· | |
| K | 322 | hCoV-19/Australia/VIC11421/2020\|EPI_ISL_563367\|202 | ···················· | ···················· | ···················· | ···················· | ···················· | ···················· | |
| K | 323 | hCoV-19/Australia/VIC11423/2020\|EPI_ISL_563368\|202 | ···················· | ···················· | ···················· | ···················· | ···················· | ···················· | |
| K | 324 | hCoV-19/Australia/VIC11431/2020\|EPI_ISL_563369\|202 | ···················· | ···················· | ···················· | ···················· | ···················· | ···················· | |
| K | 325 | hCoV-19/Australia/VIC11434/2020\|EPI_ISL_563370\|202 | ···················· | ···················· | ···················· | ···················· | ···················· | ···················· | |
| K | 326 | hCoV-19/Australia/VIC11437/2020\|EPI_ISL_563371\|202 | ···················· | ···················· | ···················· | ···················· | ···················· | ···················· | |
| K | 327 | hCoV-19/Australia/VIC11441/2020\|EPI_ISL_563372\|202 | ···················· | ···················· | ···················· | ···················· | ···················· | ···················· | |
| K | 328 | hCoV-19/Australia/VIC11442/2020\|EPI_ISL_563373\|202 | ···················· | ···················· | ···················· | ···················· | ···················· | ···················· | |
| K | 329 | hCoV-19/Australia/VIC11445/2020\|EPI_ISL_563374\|202 | ···················· | ···················· | ···················· | ···················· | ···················· | ···················· | |
| K | 330 | hCoV-19/Australia/VIC11449/2020\|EPI_ISL_563375\|202 | ···················· | ···················· | ···················· | ···················· | ···················· | ···················· | |
| K | 331 | hCoV-19/Australia/VIC11450/2020\|EPI_ISL_563376\|202 | ···················· | ···················· | ···················· | ···················· | ···················· | ···················· | |
| K | 332 | hCoV-19/Australia/VIC11451/2020\|EPI_ISL_563377\|202 | ···················· | ···················· | ···················· | ···················· | ···················· | ···················· | |
| K | 333 | hCoV-19/Australia/VIC11454/2020\|EPI_ISL_563378\|202 | ···················· | ···················· | ···················· | ···················· | ···················· | ···················· | |
| K | 334 | hCoV-19/Australia/VIC11455/2020\|EPI_ISL_563379\|202 | ···················· | ···················· | ···················· | ···················· | ···················· | ···················· | |
| K | 335 | hCoV-19/Australia/VIC11458/2020\|EPI_ISL_563380\|202 | ···················· | ···················· | ···················· | ···················· | ···················· | ···················· | |
| K | 336 | hCoV-19/Australia/VIC11459/2020\|EPI_ISL_563381\|202 | ···················· | ···················· | ···················· | ···················· | ···················· | ···················· | |
| K | 339 | hCoV-19/Australia/VIC11465/2020\|EPI_ISL_563384\|202 | ···················· | ···················· | ···················· | ···················· | ···················· | ···················· | |
| K | 340 | hCoV-19/Australia/VIC11467/2020\|EPI_ISL_563385\|202 | ···················· | ···················· | ···················· | ···················· | ···················· | ···················· | |
| K | 341 | hCoV-19/Australia/VIC11469/2020\|EPI_ISL_563386\|202 | ···················· | ···················· | ···················· | ···················· | ···················· | ···················· | |
| K | 342 | hCoV-19/Australia/VIC11470/2020\|EPI_ISL_563387\|202 | ···················· | ···················· | ···················· | ···················· | ···················· | ···················· | |
| K | 343 | hCoV-19/Australia/VIC11472/2020\|EPI_ISL_563388\|202 | ···················· | ···················· | ···················· | ···················· | ···················· | ···················· | |
| K | 344 | hCoV-19/Australia/VIC11473/2020\|EPI_ISL_563389\|202 | ···················· | ···················· | ···················· | ···················· | ···················· | ···················· | |
| K | 345 | hCoV-19/Australia/VIC11475/2020\|EPI_ISL_563390\|202 | ···················· | ···················· | ···················· | ···················· | ···················· | ···················· | |
| K | 346 | hCoV-19/Australia/VIC11476/2020\|EPI_ISL_563391\|202 | ···················· | ···················· | ···················· | ···················· | ···················· | ···················· | |
| K | 347 | hCoV-19/Australia/VIC11481/2020\|EPI_ISL_563392\|202 | ···················· | ···················· | ···················· | ···················· | ···················· | ···················· | |
| K | 349 | hCoV-19/Australia/VIC11490/2020\|EPI_ISL_563394\|202 | ···················· | ···················· | ···················· | ···················· | ···················· | ···················· | |
| K | 350 | hCoV-19/Australia/VIC11491/2020\|EPI_ISL_563395\|202 | ···················· | ···················· | ···················· | ···················· | ···················· | ···················· | |
| K | 351 | hCoV-19/Australia/VIC11495/2020\|EPI_ISL_563396\|202 | ···················· | ···················· | ···················· | ···················· | ···················· | ···················· | |
| K | 352 | hCoV-19/Australia/VIC11496/2020\|EPI_ISL_563397\|202 | ···················· | ···················· | ···················· | ···················· | ···················· | ···················· | |
| K | 353 | hCoV-19/Australia/VIC11501/2020\|EPI_ISL_563398\|202 | ···················· | ···················· | ···················· | ···················· | ···················· | ···················· | |
| K | 354 | hCoV-19/Australia/VIC11502/2020\|EPI_ISL_563399\|202 | ···················· | ···················· | ···················· | ···················· | ···················· | ···················· | |
| K | 355 | hCoV-19/Australia/VIC7717/2020\|EPI_ISL_564600\|2020 | ···················· | ···················· | ···················· | ···················· | ···················· | ···················· | |
| K | 356 | hCoV-19/Australia/VIC7718/2020\|EPI_ISL_564601\|2020 | ···················· | ···················· | ···················· | ···················· | ···················· | ···················· | |
| K | 357 | hCoV-19/Australia/VIC7719/2020\|EPI_ISL_564602\|2020 | ···················· | ···················· | ···················· | ···················· | ···················· | ···················· | |
| K | 358 | hCoV-19/Australia/VIC7720/2020\|EPI_ISL_564603\|2020 | ···················· | ···················· | ···················· | ···················· | ···················· | ···················· | |
| K | 359 | hCoV-19/Australia/VIC7721/2020\|EPI_ISL_564604\|2020 | ···················· | ···················· | ···················· | ···················· | ···················· | ···················· | |
| K | 360 | hCoV-19/Australia/VIC7722/2020\|EPI_ISL_564605\|2020 | ···················· | ···················· | ···················· | ···················· | ···················· | ···················· | |
| K | 361 | hCoV-19/Australia/VIC7723/2020\|EPI_ISL_564606\|2020 | ···················· | ···················· | ···················· | ···················· | ···················· | ···················· | |
| K | 362 | hCoV-19/Australia/VIC7724/2020\|EPI_ISL_564607\|2020 | ···················· | ···················· | ···················· | ···················· | ···················· | ···················· | |

```
K   363 hCoV-19/Australia/VIC7725/2020|EPI_ISL_564608|2020   ····················  ····················  ·····················  ····················  ····················  ····················
K   364 hCoV-19/Australia/VIC7726/2020|EPI_ISL_564609|2020   ····················  ····················  ·····················  ····················  ····················  ····················
K   365 hCoV-19/Australia/VIC7727/2020|EPI_ISL_564610|2020   ····················  ····················  ·····················  ····················  ····················  ····················
K   366 hCoV-19/Australia/VIC7728/2020|EPI_ISL_564611|2020   ····················  ····················  ·····················  ····················  ····················  ····················
K   367 hCoV-19/Australia/VIC7729/2020|EPI_ISL_564612|2020   ····················  ····················  ·····················  ····················  ····················  ····················
K   368 hCoV-19/Australia/VIC7731/2020|EPI_ISL_564613|2020   ····················  ····················  ·····················  ····················  ····················  ····················
K   369 hCoV-19/Australia/VIC7733/2020|EPI_ISL_564614|2020   ····················  ····················  ·····················  ····················  ····················  ····················
K   370 hCoV-19/Australia/VIC7734/2020|EPI_ISL_564615|2020   ····················  ····················  ·····················  ····················  ····················  ····················
K   371 hCoV-19/Australia/VIC7736/2020|EPI_ISL_564616|2020   ····················  ····················  ·····················  ····················  ····················  ····················
K   372 hCoV-19/Australia/VIC7737/2020|EPI_ISL_564617|2020   ····················  ····················  ·····················  ····················  ····················  ····················
K   373 hCoV-19/Australia/VIC7738/2020|EPI_ISL_564618|2020   ····················  ····················  ·····················  ····················  ····················  ····················
K   374 hCoV-19/Australia/VIC7741/2020|EPI_ISL_564619|2020   ····················  ····················  ·····················  ····················  ····················  ····················
K   375 hCoV-19/Australia/VIC7742/2020|EPI_ISL_564620|2020   ····················  ····················  ·····················  ····················  ····················  ····················
K   376 hCoV-19/Australia/VIC7751/2020|EPI_ISL_564629|2020   ····················  ····················  ·····················  ····················  ····················  ····················
K   377 hCoV-19/Australia/VIC7756/2020|EPI_ISL_564632|2020   ····················  ····················  ·····················  ····················  ····················  ····················
K   378 hCoV-19/Australia/VIC7758/2020|EPI_ISL_564633|2020   ····················  ····················  ·····················  ····················  ····················  ····················
K   379 hCoV-19/Australia/VIC7759/2020|EPI_ISL_564634|2020   ····················  ····················  ·····················  ····················  ····················  ····················
K   380 hCoV-19/Australia/VIC7760/2020|EPI_ISL_564635|2020   ····················  ····················  ·····················  ····················  ····················  ····················
K   381 hCoV-19/Australia/VIC7761/2020|EPI_ISL_564636|2020   ····················  ····················  ·····················  ····················  ····················  ····················
K   382 hCoV-19/Australia/VIC7762/2020|EPI_ISL_564637|2020   ····················  ····················  ·····················  ····················  ····················  ····················
K   383 hCoV-19/Australia/VIC7763/2020|EPI_ISL_564638|2020   ····················  ····················  ·····················  ····················  ····················  ····················
K   384 hCoV-19/Australia/VIC7764/2020|EPI_ISL_564639|2020   ····················  ····················  ·····················  ····················  ····················  ····················
K   385 hCoV-19/Australia/VIC7765/2020|EPI_ISL_564640|2020   ····················  ····················  ·····················  ····················  ····················  ····················
K   386 hCoV-19/Australia/VIC7766/2020|EPI_ISL_564641|2020   ····················  ····················  ·····················  ····················  ····················  ····················
K   387 hCoV-19/Australia/VIC7767/2020|EPI_ISL_564642|2020   ····················  ····················  ·····················  ····················  ····················  ····················
K   388 hCoV-19/Australia/VIC7768/2020|EPI_ISL_564643|2020   ····················  ····················  ·····················  ····················  ····················  ····················
K   389 hCoV-19/Australia/VIC7769/2020|EPI_ISL_564644|2020   ····················  ····················  ·····················  ····················  ····················  ····················
K   390 hCoV-19/Australia/VIC7772/2020|EPI_ISL_564645|2020   ····················  ····················  ·····················  ····················  ····················  ····················
K   391 hCoV-19/Australia/VIC7773/2020|EPI_ISL_564646|2020   ····················  ····················  ·····················  ····················  ····················  ····················
K   392 hCoV-19/Australia/VIC7775/2020|EPI_ISL_564647|2020   ····················  ····················  ·····················  ····················  ····················  ····················
K   393 hCoV-19/Australia/VIC7776/2020|EPI_ISL_564648|2020   ····················  ····················  ·····················  ····················  ····················  ····················
K   394 hCoV-19/Australia/VIC7777/2020|EPI_ISL_564649|2020   ····················  ····················  ·····················  ····················  ····················  ····················
K   395 hCoV-19/Australia/VIC7778/2020|EPI_ISL_564650|2020   ····················  ····················  ·····················  ····················  ····················  ····················
K   396 hCoV-19/Australia/VIC7780/2020|EPI_ISL_564651|2020   ····················  ····················  ·····················  ····················  ····················  ····················
K   397 hCoV-19/Australia/VIC7782/2020|EPI_ISL_564652|2020   ····················  ····················  ·····················  ····················  ····················  ····················
K   398 hCoV-19/Australia/VIC7783/2020|EPI_ISL_564653|2020   ····················  ····················  ·····················  ····················  ····················  ····················
K   399 hCoV-19/Australia/VIC7784/2020|EPI_ISL_564654|2020   ····················  ····················  ·····················  ····················  ····················  ····················
K   400 hCoV-19/Australia/VIC7785/2020|EPI_ISL_564655|2020   ····················  ····················  ·····················  ····················  ····················  ····················
K   401 hCoV-19/Australia/VIC7786/2020|EPI_ISL_564656|2020   ····················  ····················  ·····················  ····················  ····················  ····················
K   402 hCoV-19/Australia/VIC7787/2020|EPI_ISL_564657|2020   ····················  ····················  ·····················  ····················  ····················  ····················
K   403 hCoV-19/Australia/VIC7792/2020|EPI_ISL_564660|2020   ····················  ····················  ·····················  ····················  ····················  ····················
K   404 hCoV-19/Australia/VIC7793/2020|EPI_ISL_564661|2020   ····················  ····················  ·····················  ····················  ····················  ····················
K   405 hCoV-19/Australia/VIC7794/2020|EPI_ISL_564662|2020   ····················  ····················  ··R··················  ····················  ····················  ····················
K   406 hCoV-19/Australia/VIC7795/2020|EPI_ISL_564663|2020   ····················  ····················  ·····················  ····················  ····················  ····················
K   407 hCoV-19/Australia/VIC7796/2020|EPI_ISL_564664|2020   ····················  ····················  ·····················  ····················  ····················  ····················
K   408 hCoV-19/Australia/VIC7797/2020|EPI_ISL_564665|2020   ····················  ····················  ·····················  ····················  ····················  ····················
K   409 hCoV-19/Australia/VIC7798/2020|EPI_ISL_564666|2020   ····················  ····················  ·····················  ····················  ····················  ····················
K   410 hCoV-19/Australia/VIC7799/2020|EPI_ISL_564667|2020   ····················  ····················  ·····················  ····················  ····················  ····················
K   411 hCoV-19/Australia/VIC7803/2020|EPI_ISL_564668|2020   ····················  ····················  ·····················  ····················  ····················  ····················
K   412 hCoV-19/Australia/VIC7804/2020|EPI_ISL_564669|2020   ····················  ····················  ·····················  ····················  ····················  ····················
K   413 hCoV-19/Australia/VIC7805/2020|EPI_ISL_564670|2020   ····················  ····················  ·····················  ····················  ····················  ····················
K   414 hCoV-19/Australia/VIC7808/2020|EPI_ISL_564671|2020   ····················  ····················  ·····················  ····················  ····················  ····················
K   415 hCoV-19/Australia/VIC7811/2020|EPI_ISL_564672|2020   ····················  ····················  ·····················  ····················  ····················  ····················
K   416 hCoV-19/Australia/VIC7812/2020|EPI_ISL_564673|2020   ····················  ····················  ·····················  ····················  ····················  ····················
K   417 hCoV-19/Australia/VIC7814/2020|EPI_ISL_564674|2020   ····················  ····················  ·····················  ····················  ····················  ····················
K   418 hCoV-19/Australia/VIC7816/2020|EPI_ISL_564675|2020   ····················  ····················  ·····················  ····················  ····················  ····················
K   419 hCoV-19/Australia/VIC7818/2020|EPI_ISL_564676|2020   ····················  ····················  ·····················  ····················  ····················  ····················
K   420 hCoV-19/Australia/VIC7819/2020|EPI_ISL_564677|2020   ····················  ····················  ·····················  ····················  ····················  ····················
K   421 hCoV-19/Australia/VIC7820/2020|EPI_ISL_564678|2020   ····················  ····················  ·····················  ····················  ····················  ····················
K   422 hCoV-19/Australia/VIC7821/2020|EPI_ISL_564679|2020   ····················  ····················  ·····················  ····················  ····················  ····················
K   423 hCoV-19/Australia/VIC7824/2020|EPI_ISL_564680|2020   ····················  ····················  ·····················  ····················  ····················  ····················
K   424 hCoV-19/Australia/VIC7827/2020|EPI_ISL_564681|2020   ····················  ····················  ·····················  ····················  ····················  ····················
K   425 hCoV-19/Australia/VIC7828/2020|EPI_ISL_564682|2020   ····················  ····················  ·····················  ····················  ····················  ····················
K   426 hCoV-19/Australia/VIC7829/2020|EPI_ISL_564683|2020   ····················  ····················  ·····················  ····················  ····················  ····················
K   427 hCoV-19/Australia/VIC7830/2020|EPI_ISL_564684|2020   ····················  ····················  ·····················  ····················  ····················  ····················
K   428 hCoV-19/Australia/VIC7832/2020|EPI_ISL_564685|2020   ····················  ····················  ·····················  ····················  ····················  ····················
K   429 hCoV-19/Australia/VIC7833/2020|EPI_ISL_564686|2020   ····················  ····················  ·····················  ····················  ····················  ····················
K   430 hCoV-19/Australia/VIC7834/2020|EPI_ISL_564687|2020   ····················  ····················  ·····················  ····················  ····················  ····················
K   431 hCoV-19/Australia/VIC7835/2020|EPI_ISL_564688|2020   ····················  ····················  ·····················  ····················  ····················  ····················
K   432 hCoV-19/Australia/VIC7836/2020|EPI_ISL_564689|2020   ····················  ····················  ·····················  ····················  ····················  ····················
K   433 hCoV-19/Australia/VIC7839/2020|EPI_ISL_564690|2020   ····················  ····················  ·····················  ····················  ····················  ····················
K   434 hCoV-19/Australia/VIC7840/2020|EPI_ISL_564691|2020   ····················  ····················  ·····················  ····················  ····················  ····················
K   435 hCoV-19/Australia/VIC7841/2020|EPI_ISL_564692|2020   ····················  ····················  ·····················  ····················  ····················  ····················
K   436 hCoV-19/Australia/VIC7842/2020|EPI_ISL_564693|2020   ····················  ····················  ·····················  ····················  ····················  ····················
K   437 hCoV-19/Australia/VIC7843/2020|EPI_ISL_564694|2020   ····················  ····················  ·····················  ····················  ····················  ····················
K   438 hCoV-19/Australia/VIC7844/2020|EPI_ISL_564695|2020   ····················  ····················  ·····················  ····················  ····················  ····················
K   439 hCoV-19/Australia/VIC7845/2020|EPI_ISL_564696|2020   ····················  ····················  ·····················  ····················  ····················  ····················
K   440 hCoV-19/Australia/VIC7846/2020|EPI_ISL_564697|2020   ····················  ····················  ·····················  ····················  ····················  ····················
K   441 hCoV-19/Australia/VIC7847/2020|EPI_ISL_564698|2020   ····················  ····················  ·····················  ····················  ····················  ····················
K   442 hCoV-19/Australia/VIC7848/2020|EPI_ISL_564699|2020   ····················  ····················  ·····················  ····················  ····················  ····················
```

| | | | | | | | | |
|---|---|---|---|---|---|---|---|---|
| K | 443 | hCoV-19/USA/MI-MDHHS-SC21888/2020\|EPI_ISL_565949\|2 | ···················· | ···················· | ····················· | ···················· | ···················· | ···················· |
| K | 444 | hCoV-19/USA/MI-MDHHS-SC21890/2020\|EPI_ISL_565951\|2 | ···················· | ···················· | ····················· | ···················· | ···················· | ···················· |
| K | 445 | hCoV-19/USA/MI-MDHHS-SC21896/2020\|EPI_ISL_565957\|2 | ···················· | ···················· | ····················· | ···················· | ···················· | ···················· |
| K | 446 | hCoV-19/USA/MI-MDHHS-SC21902/2020\|EPI_ISL_565963\|2 | ···················· | ···················· | ····················· | ···················· | ···················· | ···················· |
| K | 447 | hCoV-19/USA/MI-MDHHS-SC21903/2020\|EPI_ISL_565964\|2 | ···················· | ···················· | ····················· | ···················· | ···················· | ···················· |
| K | 448 | hCoV-19/USA/MI-MDHHS-SC21904/2020\|EPI_ISL_565965\|2 | ···················· | ···················· | ····················· | ···················· | ···················· | ···················· |
| K | 449 | hCoV-19/USA/MI-MDHHS-SC21905/2020\|EPI_ISL_565966\|2 | ···················· | ···················· | ····················· | ···················· | ···················· | ···················· |
| K | 450 | hCoV-19/USA/MI-MDHHS-SC21906/2020\|EPI_ISL_565967\|2 | ···················· | ···················· | ····················· | ···················· | ···················· | ···················· |
| K | 451 | hCoV-19/USA/MI-MDHHS-SC21907/2020\|EPI_ISL_565968\|2 | ···················· | ···················· | ····················· | ···················· | ···················· | ···················· |
| K | 452 | hCoV-19/USA/MI-MDHHS-SC21908/2020\|EPI_ISL_565969\|2 | ···················· | ···················· | ····················· | ···················· | ···················· | ···················· |
| K | 453 | hCoV-19/USA/MI-MDHHS-SC21909/2020\|EPI_ISL_565970\|2 | ···················· | ···················· | ····················· | ···················· | ···················· | ···················· |
| K | 454 | hCoV-19/USA/MI-MDHHS-SC21910/2020\|EPI_ISL_565971\|2 | ···················· | ···················· | ····················· | ···················· | ···················· | ···················· |
| K | 455 | hCoV-19/USA/MI-MDHHS-SC21911/2020\|EPI_ISL_565972\|2 | ···················· | ···················· | ····················· | ···················· | ···················· | ···················· |
| K | 456 | hCoV-19/USA/MI-MDHHS-SC21912/2020\|EPI_ISL_565973\|2 | ···················· | ···················· | ····················· | ···················· | ···················· | ···················· |
| K | 457 | hCoV-19/USA/MI-MDHHS-SC21913/2020\|EPI_ISL_565974\|2 | ···················· | ···················· | ····················· | ···················· | ···················· | ···················· |
| K | 458 | hCoV-19/USA/MI-MDHHS-SC21914/2020\|EPI_ISL_565975\|2 | ···················· | ···················· | ····················· | ···················· | ···················· | ···················· |
| K | 459 | hCoV-19/USA/MI-MDHHS-SC21915/2020\|EPI_ISL_565976\|2 | ···················· | ···················· | ····················· | ···················· | ···················· | ···················· |
| K | 460 | hCoV-19/USA/MI-MDHHS-SC21916/2020\|EPI_ISL_565977\|2 | ···················· | ···················· | ····················· | ···················· | ···················· | ···················· |
| K | 461 | hCoV-19/USA/MI-MDHHS-SC21917/2020\|EPI_ISL_565978\|2 | ···················· | ···················· | ····················· | ···················· | ···················· | ···················· |
| K | 462 | hCoV-19/USA/MI-MDHHS-SC21918/2020\|EPI_ISL_565979\|2 | ···················· | ···················· | ····················· | ···················· | ···················· | ···················· |
| K | 463 | hCoV-19/USA/MI-MDHHS-SC21919/2020\|EPI_ISL_565980\|2 | ···················· | ···················· | ····················· | ···················· | ···················· | ···················· |
| K | 464 | hCoV-19/USA/MI-MDHHS-SC21921/2020\|EPI_ISL_565981\|2 | ···················· | ···················· | ····················· | ···················· | ···················· | ···················· |
| K | 465 | hCoV-19/USA/MI-MDHHS-SC21922/2020\|EPI_ISL_565982\|2 | ···················· | ···················· | ····················· | ···················· | ···················· | ···················· |
| K | 466 | hCoV-19/USA/MI-MDHHS-SC21923/2020\|EPI_ISL_565983\|2 | ···················· | ···················· | ····················· | ···················· | ···················· | ···················· |
| K | 467 | hCoV-19/USA/MI-MDHHS-SC21924/2020\|EPI_ISL_565984\|2 | ···················· | ···················· | ····················· | ···················· | ···················· | ···················· |
| K | 468 | hCoV-19/USA/MI-MDHHS-SC21925/2020\|EPI_ISL_565985\|2 | ···················· | ···················· | ····················· | ···················· | ···················· | ···················· |
| K | 469 | hCoV-19/USA/MI-MDHHS-SC21926/2020\|EPI_ISL_565986\|2 | ···················· | ···················· | ····················· | ···················· | ···················· | ···················· |
| K | 470 | hCoV-19/USA/MI-MDHHS-SC21927/2020\|EPI_ISL_565987\|2 | ···················· | ···················· | ····················· | ···················· | ···················· | ···················· |
| K | 471 | hCoV-19/USA/MI-MDHHS-SC21928/2020\|EPI_ISL_565988\|2 | ···················· | ···················· | ····················· | ···················· | ···················· | ···················· |
| K | 472 | hCoV-19/England/ALDP-9BD9A4/2020\|EPI_ISL_568171\|20 | ···················· | ···················· | ····················· | ···················· | ···················· | ···················· |
| K | 473 | hCoV-19/England/ALDP-9BEB16/2020\|EPI_ISL_568172\|20 | ···················· | ···················· | ····················· | ···················· | ···················· | ···················· |
| K | 474 | hCoV-19/England/ALDP-9BE6E8/2020\|EPI_ISL_568173\|20 | ···················· | ···················· | ····················· | ···················· | ···················· | ···················· |
| K | 475 | hCoV-19/England/ALDP-9BC7D8/2020\|EPI_ISL_568174\|20 | ···················· | ···················· | ····················· | ···················· | ···················· | ···················· |
| K | 476 | hCoV-19/England/ALDP-9BC7E7/2020\|EPI_ISL_568175\|20 | ···················· | ···················· | ····················· | ···················· | ···················· | ···················· |
| K | 477 | hCoV-19/England/ALDP-9BE4EE/2020\|EPI_ISL_568176\|20 | ···················· | ···················· | ····················· | ···················· | ···················· | ···················· |
| K | 478 | hCoV-19/England/ALDP-9BEB07/2020\|EPI_ISL_568177\|20 | ···················· | ···················· | ····················· | ···················· | ···················· | ···················· |
| K | 479 | hCoV-19/England/ALDP-9BDD8A/2020\|EPI_ISL_568178\|20 | ···················· | ···················· | ····················· | ···················· | ···················· | ···················· |
| K | 480 | hCoV-19/England/ALDP-9BDC50/2020\|EPI_ISL_568179\|20 | ···················· | ·A·················· | ····················· | ···················· | ···················· | ··A················· |
| K | 481 | hCoV-19/England/ALDP-9BC91E/2020\|EPI_ISL_568180\|20 | ···················· | ···················· | ····················· | ···················· | ···················· | ···················· |
| K | 482 | hCoV-19/England/ALDP-9BEC40/2020\|EPI_ISL_568181\|20 | ···················· | ···················· | ····················· | ···················· | ···················· | ···················· |
| K | 483 | hCoV-19/England/ALDP-9BC8A8/2020\|EPI_ISL_568183\|20 | ···················· | ···················· | ····················· | ···················· | ···················· | ···················· |
| K | 484 | hCoV-19/England/ALDP-9BE28A/2020\|EPI_ISL_568184\|20 | ···················· | ···················· | ····················· | ···················· | ···················· | ···················· |
| K | 485 | hCoV-19/England/ALDP-9BECF5/2020\|EPI_ISL_568185\|20 | ···················· | ···················· | ····················· | ···················· | ···················· | ···················· |
| K | 486 | hCoV-19/England/ALDP-9BE703/2020\|EPI_ISL_568186\|20 | ···················· | ···················· | ····················· | ···················· | ···················· | ···················· |
| K | 487 | hCoV-19/England/ALDP-9BC6F9/2020\|EPI_ISL_568188\|20 | ···················· | ···················· | ····················· | ···················· | ···················· | ···················· |
| K | 488 | hCoV-19/England/ALDP-9BDBF9/2020\|EPI_ISL_568189\|20 | ···················· | ···················· | ····················· | ···················· | ···················· | ···················· |
| K | 489 | hCoV-19/England/ALDP-9BE7F4/2020\|EPI_ISL_568190\|20 | ···················· | ···················· | ····················· | ···················· | ···················· | ···················· |
| K | 490 | hCoV-19/England/ALDP-9BDAA1/2020\|EPI_ISL_568191\|20 | ···················· | ···················· | ····················· | ···················· | ···················· | ···················· |
| K | 491 | hCoV-19/England/ALDP-9BDB17/2020\|EPI_ISL_568193\|20 | ···················· | ···················· | ····················· | ···················· | ···················· | ···················· |
| K | 492 | hCoV-19/England/ALDP-9BDAB0/2020\|EPI_ISL_568194\|20 | ···················· | ···················· | ····················· | ···················· | ···················· | ···················· |
| K | 493 | hCoV-19/England/ALDP-9BE41B/2020\|EPI_ISL_568195\|20 | ···················· | ···················· | ····················· | ···················· | ···················· | ···················· |
| K | 494 | hCoV-19/England/ALDP-9BDD7B/2020\|EPI_ISL_568196\|20 | ···················· | ···················· | ····················· | ···················· | ···················· | ···················· |
| K | 495 | hCoV-19/England/ALDP-9BE509/2020\|EPI_ISL_568197\|20 | ···················· | ···················· | ····················· | ···················· | ···················· | ···················· |
| K | 496 | hCoV-19/England/ALDP-9BC76F/2020\|EPI_ISL_568198\|20 | ···················· | ···················· | ····················· | ···················· | ···················· | ···················· |
| K | 497 | hCoV-19/England/ALDP-9BE09F/2020\|EPI_ISL_568199\|20 | ···················· | ···················· | ····················· | ···················· | ···················· | ···················· |
| K | 498 | hCoV-19/USA/WA-UW-12070/2020\|EPI_ISL_570484\|2020-0 | ···················· | ···················· | ····················· | ···················· | ···················· | ···················· |
| K | 499 | hCoV-19/USA/WA-UW-12078/2020\|EPI_ISL_570485\|2020-0 | ···················· | ·A·················· | ····················· | ···················· | ···················· | ···················· |
| K | 500 | hCoV-19/USA/WA-UW-12085/2020\|EPI_ISL_570486\|2020-0 | ···················· | ···················· | ····················· | ···················· | ···················· | ···················· |
| K | 501 | hCoV-19/USA/OR-UW-12088/2020\|EPI_ISL_570487\|2020-0 | ···················· | ···················· | ····················· | ···················· | ···················· | ···················· |
| K | 502 | hCoV-19/USA/WA-UW-12097/2020\|EPI_ISL_570488\|2020-0 | ···················· | ···················· | ····················· | ···················· | ···················· | ···················· |
| K | 503 | hCoV-19/USA/WA-UW-12098/2020\|EPI_ISL_570489\|2020-0 | ···················· | ···················· | ····················· | ···················· | ···················· | ···················· |
| K | 504 | hCoV-19/USA/WA-UW-12117/2020\|EPI_ISL_570490\|2020-0 | ···················· | ···················· | ····················· | ···················· | ···················· | ···················· |
| K | 505 | hCoV-19/USA/WA-UW-12127/2020\|EPI_ISL_570491\|2020-0 | ···················· | ···················· | ····················· | ···················· | ···················· | ···················· |
| K | 506 | hCoV-19/USA/WA-UW-12134/2020\|EPI_ISL_570492\|2020-0 | ···················· | ·A·················· | ····················· | ···················· | ···················· | ···················· |
| K | 507 | hCoV-19/USA/WA-UW-12140/2020\|EPI_ISL_570493\|2020-0 | ···················· | ·A·················· | ····················· | ···················· | ···················· | ···················· |
| K | 508 | hCoV-19/USA/WA-UW-12141/2020\|EPI_ISL_570494\|2020-0 | ···················· | ·A·················· | ····················· | ···················· | ···················· | ···················· |
| K | 509 | hCoV-19/USA/WA-UW-12162/2020\|EPI_ISL_570496\|2020-0 | ···················· | ···················· | ····················· | ···················· | ···················· | ···················· |
| K | 510 | hCoV-19/USA/WA-UW-12163/2020\|EPI_ISL_570497\|2020-0 | ···················· | ···················· | ····················· | ···················· | ···················· | ···················· |
| K | 511 | hCoV-19/USA/WA-UW-12164/2020\|EPI_ISL_570498\|2020-0 | ···················· | ···················· | ····················· | ···················· | ···················· | ···················· |
| K | 512 | hCoV-19/USA/WA-UW-12167/2020\|EPI_ISL_570499\|2020-0 | ···················· | ···················· | ····················· | ···················· | ···················· | ···················· |
| K | 513 | hCoV-19/USA/TX-DSHS-0517/2020\|EPI_ISL_560960\|2020- | ···················· | ···················· | ····················· | ···················· | ···················· | ···················· |
| K | 514 | hCoV-19/USA/TX-DSHS-0520/2020\|EPI_ISL_560962\|2020- | ···················· | ···················· | ····················· | ···················· | ···················· | ···················· |
| K | 515 | hCoV-19/USA/TX-DSHS-0523/2020\|EPI_ISL_560965\|2020- | ···················· | ···················· | ····················· | ···················· | ···················· | ···················· |
| K | 516 | hCoV-19/USA/TX-DSHS-0524/2020\|EPI_ISL_560966\|2020- | ···················· | ···················· | ····················· | ···················· | ···················· | ···················· |
| K | 517 | hCoV-19/USA/TX-DSHS-0525/2020\|EPI_ISL_560967\|2020- | ···················· | ···················· | ····················· | ···················· | ···················· | ···················· |
| K | 518 | hCoV-19/USA/TX-DSHS-0526/2020\|EPI_ISL_560968\|2020- | ···················· | ···················· | ····················· | ···················· | ···················· | ···················· |
| K | 519 | hCoV-19/USA/TX-DSHS-0527/2020\|EPI_ISL_560969\|2020- | ···················· | ···················· | ····················· | ···················· | ···················· | ···················· |
| K | 520 | hCoV-19/Sweden/20-50303/2020\|EPI_ISL_560972\|2020-0 | ···················· | ···················· | ····················· | ···················· | ···················· | ···················· |
| K | 521 | hCoV-19/Sweden/20-51749/2020\|EPI_ISL_560973\|2020-0 | ···················· | ···················· | ····················· | ···················· | ···················· | ···················· |
| K | 522 | hCoV-19/Sweden/20-51952/2020\|EPI_ISL_560974\|2020-0 | ···················· | ···················· | ····················· | ···················· | ···················· | ···················· |

| | | | | | | | |
|---|---|---|---|---|---|---|---|
| K | 523 hCoV-19/Sweden/20-52037/2020\|EPI_ISL_560975\|2020-0 | .................... | .................... | .................... | .................... | .................... | .................... |
| K | 524 hCoV-19/Sweden/20-52076/2020\|EPI_ISL_560976\|2020-0 | .................... | .................... | .................... | .................... | .................... | .................... |
| K | 525 hCoV-19/Sweden/20-08929/2020\|EPI_ISL_560977\|2020-0 | .................... | .................... | .................... | .................... | .................... | .................... |
| K | 526 hCoV-19/Sweden/20-08930/2020\|EPI_ISL_560978\|2020-0 | .................... | .................... | .................... | .................... | .................... | .................... |
| K | 527 hCoV-19/Sweden/20-08931/2020\|EPI_ISL_560979\|2020-0 | .................... | .................... | .................... | .................... | .................... | .................... |
| K | 528 hCoV-19/Sweden/20-08936/2020\|EPI_ISL_560980\|2020-0 | .................... | .................... | .................... | .................... | .................... | .................... |
| K | 529 hCoV-19/Sweden/20-08946/2020\|EPI_ISL_560981\|2020-0 | .................... | .................... | .................... | .................... | .................... | .................... |
| K | 530 hCoV-19/Sweden/20-52279/2020\|EPI_ISL_560982\|2020-0 | .................... | .................... | .................... | .................... | .................... | .................... |
| K | 531 hCoV-19/Sweden/20-52280/2020\|EPI_ISL_560983\|2020-0 | .................... | .................... | .................... | .................... | .................... | .................... |
| K | 532 hCoV-19/Sweden/20-52281/2020\|EPI_ISL_560984\|2020-0 | .................... | .................... | .................... | .................... | .................... | .................... |
| K | 533 hCoV-19/Sweden/20-52285/2020\|EPI_ISL_560985\|2020-0 | .................... | .................... | .................... | .................... | .................... | .................... |
| K | 534 hCoV-19/Sweden/20-52419/2020\|EPI_ISL_560986\|2020-0 | .................... | .................... | .................... | .................... | .................... | .................... |
| K | 535 hCoV-19/Sweden/20-52426/2020\|EPI_ISL_560987\|2020-0 | .................... | .................... | .................... | .................... | .................... | .................... |
| K | 536 hCoV-19/Sweden/20-52586/2020\|EPI_ISL_560988\|2020-0 | .................... | .................... | .................... | .................... | .................... | .................... |
| K | 537 hCoV-19/Sweden/20-52646/2020\|EPI_ISL_560989\|2020-0 | .................... | .................... | .................... | .................... | .................... | .................... |
| K | 538 hCoV-19/Sweden/20-52649/2020\|EPI_ISL_560990\|2020-0 | .................... | .................... | .................... | .................... | .................... | .................... |
| K | 539 hCoV-19/Indonesia/EJ-ITD-3601NT/2020\|EPI_ISL_56099 | .................... | .................... | .................... | .................... | .................... | .................... |
| K | 543 hCoV-19/Gambia/3678/2020\|EPI_ISL_560997\|2020-07-30 | .................... | .................... | .................... | .................... | .................... | .................... |
| K | 544 hCoV-19/Australia/VIC10931/2020\|EPI_ISL_563158\|202 | .................... | .................... | .................... | .................... | .................... | .................... |
| K | 545 hCoV-19/Australia/VIC10932/2020\|EPI_ISL_563159\|202 | .................... | .................... | .................... | .................... | .................... | .................... |
| K | 546 hCoV-19/Australia/VIC10944/2020\|EPI_ISL_563161\|202 | .................... | .................... | .................... | .................... | .................... | .................... |
| K | 547 hCoV-19/Australia/VIC10947/2020\|EPI_ISL_563162\|202 | .................... | .................... | .................... | .................... | .................... | .................... |
| K | 548 hCoV-19/Australia/VIC10948/2020\|EPI_ISL_563163\|202 | .................... | .................... | .................... | .................... | .................... | .................... |
| K | 549 hCoV-19/Australia/VIC10954/2020\|EPI_ISL_563164\|202 | .................... | .................... | .................... | .................... | .................... | .................... |
| K | 550 hCoV-19/Australia/VIC10955/2020\|EPI_ISL_563165\|202 | .................... | .................... | .................... | .................... | .................... | .................... |
| K | 551 hCoV-19/Australia/VIC10956/2020\|EPI_ISL_563166\|202 | .................... | .................... | .................... | .................... | .................... | .................... |
| K | 552 hCoV-19/Australia/VIC10957/2020\|EPI_ISL_563167\|202 | .................... | .................... | .................... | .................... | N................... | .................... |
| K | 553 hCoV-19/Australia/VIC10958/2020\|EPI_ISL_563168\|202 | .................... | .................... | .................... | .................... | .................... | .................... |
| K | 554 hCoV-19/Australia/VIC10959/2020\|EPI_ISL_563169\|202 | .................... | .................... | .................... | .................... | .................... | .................... |
| K | 555 hCoV-19/Australia/VIC10961/2020\|EPI_ISL_563170\|202 | .................... | .................... | .................... | .................... | .................... | .................... |
| K | 556 hCoV-19/Australia/VIC10963/2020\|EPI_ISL_563171\|202 | .................... | .................... | .................... | .................... | .................... | .................... |
| K | 557 hCoV-19/Australia/VIC10965/2020\|EPI_ISL_563172\|202 | .................... | .................... | .................... | .................... | .................... | .................... |
| K | 558 hCoV-19/Australia/VIC10967/2020\|EPI_ISL_563173\|202 | .................... | .................... | .................... | .................... | .................... | .................... |
| K | 559 hCoV-19/Australia/VIC10968/2020\|EPI_ISL_563174\|202 | .................... | .................... | .................... | .................... | .................... | .................... |
| K | 560 hCoV-19/Australia/VIC10969/2020\|EPI_ISL_563175\|202 | .................... | .................... | .................... | .................... | .................... | .................... |
| K | 561 hCoV-19/Australia/VIC10970/2020\|EPI_ISL_563176\|202 | .................... | .................... | .................... | .................... | .................... | .................... |
| K | 562 hCoV-19/Australia/VIC10973/2020\|EPI_ISL_563177\|202 | .................... | .................... | .................... | .................... | .................... | .................... |
| K | 564 hCoV-19/Australia/VIC10980/2020\|EPI_ISL_563179\|202 | .................... | .................... | .................... | .................... | .................... | .................... |
| K | 566 hCoV-19/Australia/VIC7546/2020\|EPI_ISL_564489\|2020 | .................... | .................... | .................... | .................... | .................... | .................... |
| K | 567 hCoV-19/Australia/VIC7547/2020\|EPI_ISL_564490\|2020 | .................... | .................... | .................... | .................... | .................... | .................... |
| K | 568 hCoV-19/Australia/VIC7550/2020\|EPI_ISL_564492\|2020 | .................... | .................... | .................... | .................... | .................... | .................... |
| K | 569 hCoV-19/Australia/VIC7551/2020\|EPI_ISL_564493\|2020 | .................... | .................... | .................... | .................... | .................... | .................... |
| K | 570 hCoV-19/Australia/VIC7553/2020\|EPI_ISL_564494\|2020 | .................... | .................... | .................... | .................... | .................... | .................... |
| K | 571 hCoV-19/Australia/VIC7555/2020\|EPI_ISL_564495\|2020 | .................... | .................... | .................... | .................... | .................... | .................... |
| K | 572 hCoV-19/Australia/VIC7556/2020\|EPI_ISL_564496\|2020 | .................... | .................... | .................... | .................... | .................... | .................... |
| K | 573 hCoV-19/Australia/VIC7558/2020\|EPI_ISL_564497\|2020 | .................... | .................... | .................... | .................... | .................... | .................... |
| K | 574 hCoV-19/Australia/VIC7560/2020\|EPI_ISL_564498\|2020 | .................... | .................... | .................... | .................... | .................... | .................... |
| K | 575 hCoV-19/Australia/VIC7561/2020\|EPI_ISL_564499\|2020 | .................... | .................... | .................... | .................... | .................... | .................... |
| K | 576 hCoV-19/Australia/VIC7743/2020\|EPI_ISL_564621\|2020 | .................... | .................... | .................... | .................... | .................... | .................... |
| K | 577 hCoV-19/Australia/VIC7744/2020\|EPI_ISL_564622\|2020 | .................... | .................... | .................... | .................... | .................... | .................... |
| K | 578 hCoV-19/Australia/VIC7745/2020\|EPI_ISL_564623\|2020 | .................... | .................... | .................... | .................... | .................... | .................... |
| K | 579 hCoV-19/Australia/VIC7746/2020\|EPI_ISL_564624\|2020 | .................... | .................... | .................... | .................... | .................... | .................... |
| K | 580 hCoV-19/Australia/VIC7747/2020\|EPI_ISL_564625\|2020 | .................... | .................... | .................... | .................... | .................... | .................... |
| K | 581 hCoV-19/Australia/VIC7748/2020\|EPI_ISL_564626\|2020 | .................... | .................... | .................... | .................... | .................... | .................... |
| K | 582 hCoV-19/Australia/VIC7749/2020\|EPI_ISL_564627\|2020 | .................... | .................... | .................... | .................... | .................... | .................... |
| K | 583 hCoV-19/Australia/VIC7750/2020\|EPI_ISL_564628\|2020 | .................... | .................... | .................... | .................... | .................... | .................... |
| K | 584 hCoV-19/Australia/VIC7754/2020\|EPI_ISL_564630\|2020 | .................... | .................... | .................... | .................... | .................... | .................... |
| K | 585 hCoV-19/Australia/VIC7755/2020\|EPI_ISL_564631\|2020 | .................... | .................... | .................... | .................... | .................... | .................... |
| K | 586 hCoV-19/USA/MI-MDHHS-SC21892/2020\|EPI_ISL_565953\|2 | .................... | .................... | .................... | .................... | .................... | .................... |
| K | 587 hCoV-19/USA/MI-MDHHS-SC21893/2020\|EPI_ISL_565954\|2 | .................... | .................... | .................... | .................... | .................... | .................... |
| K | 588 hCoV-19/USA/MI-MDHHS-SC21894/2020\|EPI_ISL_565955\|2 | .................... | .................... | .................... | .................... | .................... | .................... |
| K | 589 hCoV-19/USA/MI-MDHHS-SC21895/2020\|EPI_ISL_565956\|2 | .................... | .................... | .................... | .................... | .................... | .................... |
| K | 590 hCoV-19/USA/MI-MDHHS-SC21897/2020\|EPI_ISL_565958\|2 | .................... | .................... | .................... | .................... | .................... | .................... |
| K | 591 hCoV-19/USA/MI-MDHHS-SC21898/2020\|EPI_ISL_565959\|2 | .................... | .................... | .................... | .................... | .................... | .................... |
| K | 592 hCoV-19/USA/MI-MDHHS-SC21899/2020\|EPI_ISL_565960\|2 | .................... | .................... | .................... | .................... | .................... | .................... |
| K | 593 hCoV-19/USA/MI-MDHHS-SC21900/2020\|EPI_ISL_565961\|2 | .................... | .................... | .................... | .................... | .................... | .................... |
| K | 594 hCoV-19/USA/MI-MDHHS-SC21901/2020\|EPI_ISL_565962\|2 | .................... | .................... | .................... | .................... | .................... | .................... |
| K | 595 hCoV-19/England/ALDP-9BE536/2020\|EPI_ISL_568000\|20 | .................... | .................... | .................... | .................... | .................... | .................... |
| K | 596 hCoV-19/England/ALDP-9BDC23/2020\|EPI_ISL_568001\|20 | .................... | .................... | .................... | .................... | .................... | .................... |
| K | 597 hCoV-19/England/ALDP-9BC69F/2020\|EPI_ISL_568002\|20 | .................... | .................... | .................... | .................... | .................... | .................... |
| K | 598 hCoV-19/England/ALDP-9BE68E/2020\|EPI_ISL_568003\|20 | .................... | .................... | .................... | .................... | .................... | .................... |
| K | 599 hCoV-19/England/ALDP-9BED89/2020\|EPI_ISL_568004\|20 | .................... | .................... | .................... | .................... | .................... | .................... |
| K | 600 hCoV-19/England/ALDP-9BE651/2020\|EPI_ISL_568005\|20 | .................... | .................... | .................... | .................... | .................... | .................... |
| K | 601 hCoV-19/England/ALDP-9BEA91/2020\|EPI_ISL_568006\|20 | .................... | .................... | .................... | .................... | .................... | .................... |
| K | 602 hCoV-19/England/ALDP-9BE6CA/2020\|EPI_ISL_568007\|20 | .................... | .................... | .................... | .................... | .................... | .................... |
| K | 603 hCoV-19/England/ALDP-9BDC6F/2020\|EPI_ISL_568008\|20 | .................... | .................... | .................... | .................... | .................... | .................... |
| K | 604 hCoV-19/England/ALDP-9BDCD8/2020\|EPI_ISL_568009\|20 | .................... | .................... | .................... | .................... | .................... | .................... |
| K | 605 hCoV-19/England/ALDP-9BE985/2020\|EPI_ISL_568010\|20 | .................... | .................... | .................... | .................... | .................... | .................... |
| K | 606 hCoV-19/England/ALDP-9BE40C/2020\|EPI_ISL_568011\|20 | .................... | .................... | .................... | .................... | .................... | .................... |
| K | 607 hCoV-19/England/ALDP-9BE67F/2020\|EPI_ISL_568013\|20 | .................... | .................... | .................... | .................... | .................... | .................... |

K 608 hCoV-19/England/ALDP-9BEC22/2020|EPI_ISL_568014|20 ···················· ···················· ····················· ···················· ···················· ····················
K 609 hCoV-19/England/ALDP-9BE1F6/2020|EPI_ISL_568016|20 ···················· ···················· ····················· ···················· ···················· ····················
K 610 hCoV-19/England/ALDP-9BE9B2/2020|EPI_ISL_568017|20 ···················· ···················· ····················· ···················· ···················· ····················
K 611 hCoV-19/England/ALDP-9BEA0A/2020|EPI_ISL_568018|20 ···················· ···················· ····················· ···················· ···················· ····················
K 612 hCoV-19/England/ALDP-9BD9C2/2020|EPI_ISL_568019|20 ···················· ···················· ····················· ···················· ···················· ····················
K 613 hCoV-19/England/ALDP-9BEDD4/2020|EPI_ISL_568020|20 ···················· ·A·······A·········· ····················· ···················· ···················· ····················
K 614 hCoV-19/England/ALDP-9BEB70/2020|EPI_ISL_568021|20 ···················· ·A·················· ····················· ···················· ···················· ···A················
K 615 hCoV-19/England/ALDP-9BC723/2020|EPI_ISL_568022|20 ···················· ···················· ····················· ···················· ···················· ····················
K 616 hCoV-19/England/ALDP-9BDE96/2020|EPI_ISL_568024|20 ···················· ···················· ····················· ···················· ···················· ····················
K 617 hCoV-19/England/ALDP-9BE071/2020|EPI_ISL_568025|20 ···················· ···················· ····················· ···················· ···················· ····················
K 618 hCoV-19/England/ALDP-9BDEF0/2020|EPI_ISL_568027|20 ···················· ···················· ····················· ···················· ···················· ····················
K 619 hCoV-19/England/ALDP-9BE7E5/2020|EPI_ISL_568030|20 ···················· ···················· ····················· ···················· ···················· ····················
K 620 hCoV-19/England/ALDP-9BE9C1/2020|EPI_ISL_568031|20 ···················· ···················· ····················· ···················· ···················· ····················
K 621 hCoV-19/England/ALDP-9BE114/2020|EPI_ISL_568033|20 ···················· ···················· ····················· ···················· ···················· ····················
K 622 hCoV-19/England/ALDP-9BC84E/2020|EPI_ISL_568034|20 ···················· ···················· ····················· ···················· ···················· ····················
K 623 hCoV-19/England/ALDP-9BC811/2020|EPI_ISL_568035|20 ···················· ···················· ····················· ···················· ···················· ····················
K 624 hCoV-19/England/ALDP-9BEAEC/2020|EPI_ISL_568036|20 ···················· ···················· ····················· ···················· ···················· ····················
K 625 hCoV-19/England/ALDP-9BDDA8/2020|EPI_ISL_568038|20 ···················· ···················· ····················· ···················· ···················· ····················
K 626 hCoV-19/England/ALDP-9BDCF6/2020|EPI_ISL_568039|20 ···················· ···················· ····················· ···················· ···················· ····················
K 627 hCoV-19/England/ALDP-9BE053/2020|EPI_ISL_568041|20 ···················· ···················· ····················· ···················· ···················· ····················
K 628 hCoV-19/England/ALDP-9BEB8F/2020|EPI_ISL_568042|20 ···················· ···················· ····················· ···················· ···················· ····················
K 629 hCoV-19/England/ALDP-9BEC7D/2020|EPI_ISL_568043|20 ···················· ···················· ····················· ···················· ···················· ····················
K 630 hCoV-19/England/ALDP-9BDA38/2020|EPI_ISL_568044|20 ···················· ···················· ····················· ···················· ···················· ····················
K 631 hCoV-19/England/ALDP-9BE2B7/2020|EPI_ISL_568045|20 ···················· ·A·················· ····················· ···················· ···················· ····················
K 632 hCoV-19/England/ALDP-9BEABF/2020|EPI_ISL_568046|20 ···················· ···················· ····················· ···················· ···················· ····················
K 633 hCoV-19/England/ALDP-9BD9D1/2020|EPI_ISL_568047|20 ···················· ·A·················· ····················· ···················· ···················· ····················
K 634 hCoV-19/England/ALDP-9BDE69/2020|EPI_ISL_568048|20 ···················· ···················· ····················· ···················· ···················· ····················
K 635 hCoV-19/England/ALDP-9BE25D/2020|EPI_ISL_568049|20 ···················· ···················· ····················· ···················· ···················· ····················
K 636 hCoV-19/France/PAC-IHU-0084/2020|EPI_ISL_569300|20 ···················· ···················· ····················· ···················· ···················· ····················
K 637 hCoV-19/France/PAC-IHU-0086/2020|EPI_ISL_569301|20 ···················· ···················· ····················· ···················· ···················· ····················
K 638 hCoV-19/France/PAC-IHU-0087/2020|EPI_ISL_569302|20 ···················· ···················· ····················· ···················· ···················· ····················
K 639 hCoV-19/France/PAC-IHU-0092/2020|EPI_ISL_569303|20 ···················· ···················· ····················· ···················· ···················· ····················
K 640 hCoV-19/France/PAC-IHU-0093/2020|EPI_ISL_569304|20 ···················· ···················· ····················· ···················· ···················· ····················
K 641 hCoV-19/France/PAC-IHU-0094/2020|EPI_ISL_569305|20 ···················· ···················· ····················· ···················· ···················· ····················
K 642 hCoV-19/France/PAC-IHU-0097/2020|EPI_ISL_569307|20 ···················· ···················· ····················· ···················· ···················· ····················
K 643 hCoV-19/France/PAC-IHU-0099/2020|EPI_ISL_569308|20 ···················· ···················· ····················· ···················· ···················· ····················
K 644 hCoV-19/France/PAC-IHU-0100/2020|EPI_ISL_569309|20 ···················· ···················· ····················· ···················· ···················· ····················
K 645 hCoV-19/France/PAC-IHU-0104/2020|EPI_ISL_569310|20 ···················· ···················· ····················· ···················· ···················· ····················
K 646 hCoV-19/France/PAC-IHU-0106/2020|EPI_ISL_569311|20 ···················· ···················· ····················· ···················· ···················· ····················
K 647 hCoV-19/France/PAC-IHU-0107/2020|EPI_ISL_569312|20 ···················· ···················· ····················· ···················· ···················· ····················
K 648 hCoV-19/France/PAC-IHU-0108/2020|EPI_ISL_569313|20 ···················· ···················· ····················· ···················· ···················· ····················
K 649 hCoV-19/France/PAC-IHU-0109/2020|EPI_ISL_569314|20 ···················· ···················· ····················· ···················· ···················· ····················
K 650 hCoV-19/France/PAC-IHU-0115/2020|EPI_ISL_569315|20 ···················· ···················· ····················· ···················· ···················· ····················
K 651 hCoV-19/France/PAC-IHU-0116/2020|EPI_ISL_569316|20 ···················· ···················· ····················· ···················· ···················· ····················
K 652 hCoV-19/France/PAC-IHU-0117/2020|EPI_ISL_569317|20 ···················· ···················· ····················· ···················· ···················· ····················
K 653 hCoV-19/France/PAC-IHU-0118/2020|EPI_ISL_569318|20 ···················· ···················· ····················· ···················· ···················· ····················
K 654 hCoV-19/France/PAC-IHU-0122/2020|EPI_ISL_569321|20 ···················· ···················· ····················· ···················· ···················· ····················
K 655 hCoV-19/France/PAC-IHU-0126/2020|EPI_ISL_569323|20 ···················· ···················· ····················· ···················· ···················· ····················
K 656 hCoV-19/France/PAC-IHU-0128/2020|EPI_ISL_569324|20 ···················· ···················· ····················· ···················· ···················· ····················
K 657 hCoV-19/France/PAC-IHU-0129/2020|EPI_ISL_569325|20 ···················· ···················· ····················· ···················· ···················· ····················
K 658 hCoV-19/France/PAC-IHU-0130/2020|EPI_ISL_569326|20 ···················· ···················· ····················· ···················· ···················· ····················
K 659 hCoV-19/France/PAC-IHU-0131/2020|EPI_ISL_569327|20 ···················· ···················· ····················· ···················· ···················· ····················
K 660 hCoV-19/France/PAC-IHU-0132/2020|EPI_ISL_569328|20 ···················· ···················· ····················· ···················· ···················· ····················
K 661 hCoV-19/France/PAC-IHU-0133/2020|EPI_ISL_569329|20 ···················· ···················· ····················· ···················· ···················· ····················
K 662 hCoV-19/France/PAC-IHU-0135/2020|EPI_ISL_569330|20 ···················· ···················· ····················· ···················· ···················· ····················
K 663 hCoV-19/France/PAC-IHU-0136/2020|EPI_ISL_569331|20 ···················· ···················· ····················· ···················· ···················· ····················
K 664 hCoV-19/France/PAC-IHU-0139/2020|EPI_ISL_569332|20 ···················· ···················· ····················· ···················· ···················· ····················
K 665 hCoV-19/France/PAC-IHU-0141/2020|EPI_ISL_569333|20 ···················· ···················· ····················· ···················· ···················· ····················
K 666 hCoV-19/France/PAC-IHU-0145/2020|EPI_ISL_569334|20 ···················· ···················· ····················· ···················· ···················· ····················
K 667 hCoV-19/France/PAC-IHU-0146/2020|EPI_ISL_569335|20 ···················· ···················· ····················· ···················· ···················· ····················
K 668 hCoV-19/France/PAC-IHU-0149/2020|EPI_ISL_569336|20 ···················· ···················· ····················· ···················· ···················· ····················
K 669 hCoV-19/France/PAC-IHU-0151/2020|EPI_ISL_569337|20 ···················· ···················· ····················· ···················· ···················· ····················
K 670 hCoV-19/France/PAC-IHU-0152/2020|EPI_ISL_569338|20 ···················· ···················· ····················· ···················· ···················· ····················
K 671 hCoV-19/France/PAC-IHU-0153/2020|EPI_ISL_569339|20 ···················· ···················· ····················· ···················· ···················· ····················
K 672 hCoV-19/France/PAC-IHU-0155/2020|EPI_ISL_569340|20 ···················· ··········A········· ····················· ···················· ···················· ····················
K 673 hCoV-19/France/PAC-IHU-0158/2020|EPI_ISL_569341|20 ···················· ···················· ····················· ···················· ···················· ····················
K 674 hCoV-19/France/PAC-IHU-0162/2020|EPI_ISL_569343|20 ···················· ···················· ····················· ···················· ···················· ····················
K 675 hCoV-19/France/PAC-IHU-0167/2020|EPI_ISL_569344|20 ···················· ···················· ····················· ···················· ···················· ····················
K 676 hCoV-19/France/PAC-IHU-0168/2020|EPI_ISL_569345|20 ···················· ···················· ····················· ···················· ···················· ····················
K 677 hCoV-19/France/PAC-IHU-0170/2020|EPI_ISL_569346|20 ···················· ···················· ····················· ···················· ···················· ····················
K 678 hCoV-19/France/PAC-IHU-0171/2020|EPI_ISL_569347|20 ···················· ···················· ····················· ···················· ···················· ····················
K 679 hCoV-19/France/PAC-IHU-0172/2020|EPI_ISL_569348|20 ···················· ···················· ····················· ···················· ···················· ····················
K 680 hCoV-19/France/PAC-IHU-0177/2020|EPI_ISL_569350|20 ···················· ···················· ····················· ···················· ···················· ····················
K 681 hCoV-19/France/PAC-IHU-0178/2020|EPI_ISL_569351|20 ···················· ···················· ····················· ···················· ···················· ····················
K 682 hCoV-19/France/PAC-IHU-0179/2020|EPI_ISL_569352|20 ···················· ···················· ····················· ···················· ···················· ····················
K 683 hCoV-19/France/PAC-IHU-0180/2020|EPI_ISL_569353|20 ···················· ···················· ····················· ···················· ···················· ····················
K 684 hCoV-19/France/PAC-IHU-0186/2020|EPI_ISL_569354|20 ···················· ···················· ····················· ···················· ···················· ····················
K 685 hCoV-19/France/PAC-IHU-0187/2020|EPI_ISL_569355|20 ···················· ···················· ····················· ···················· ···················· ····················
K 686 hCoV-19/France/PAC-IHU-0188/2020|EPI_ISL_569356|20 ···················· ··········A········· ····················· ···················· ···················· ····················
K 687 hCoV-19/France/PAC-IHU-0189/2020|EPI_ISL_569357|20 ···················· ···················· ····················· ···················· ···················· ····················

| | | | | | | | | |
|---|---|---|---|---|---|---|---|---|
| K | 688 | hCoV-19/France/PAC-IHU-0190/2020\|EPI_ISL_569358\|20 | ···················· | ···················· | ····················· | ···················· | ···················· | ···················· |
| K | 689 | hCoV-19/France/PAC-IHU-0191/2020\|EPI_ISL_569359\|20 | ···················· | ···················· | ····················· | ···················· | ···················· | ···················· |
| K | 690 | hCoV-19/France/PAC-IHU-0193/2020\|EPI_ISL_569360\|20 | ···················· | ···················· | ····················· | ···················· | ···················· | ···················· |
| K | 691 | hCoV-19/France/PAC-IHU-0194/2020\|EPI_ISL_569361\|20 | ···················· | ···················· | ····················· | ···················· | ···················· | ···················· |
| K | 692 | hCoV-19/France/PAC-IHU-0195/2020\|EPI_ISL_569362\|20 | ···················· | ···················· | ····················· | ···················· | ···················· | ···················· |
| K | 693 | hCoV-19/France/PAC-IHU-0196/2020\|EPI_ISL_569363\|20 | ···················· | ···················· | ····················· | ···················· | ···················· | ···················· |
| K | 694 | hCoV-19/France/PAC-IHU-0197/2020\|EPI_ISL_569364\|20 | ···················· | ···················· | ····················· | ···················· | ···················· | ···················· |
| K | 695 | hCoV-19/France/PAC-IHU-0198/2020\|EPI_ISL_569365\|20 | ···················· | ···················· | ····················· | ···················· | ···················· | ···················· |
| K | 696 | hCoV-19/France/PAC-IHU-0199/2020\|EPI_ISL_569366\|20 | ···················· | ···················· | ····················· | ···················· | ···················· | ···················· |
| K | 697 | hCoV-19/France/PAC-IHU-0200/2020\|EPI_ISL_569367\|20 | ···················· | ···················· | ····················· | ···················· | ···················· | ···················· |
| K | 698 | hCoV-19/France/PAC-IHU-0201/2020\|EPI_ISL_569368\|20 | ···················· | ···················· | ····················· | ···················· | ···················· | ···················· |
| K | 699 | hCoV-19/France/PAC-IHU-0203/2020\|EPI_ISL_569369\|20 | ···················· | ····T··············· | ····················· | ···················· | ···················· | ···················· |
| K | 700 | hCoV-19/France/PAC-IHU-0204/2020\|EPI_ISL_569370\|20 | ···················· | ···················· | ····················· | ···················· | ···················· | ···················· |
| K | 701 | hCoV-19/France/PAC-IHU-0205/2020\|EPI_ISL_569371\|20 | ···················· | ···················· | ····················· | ···················· | ···················· | ···················· |
| K | 702 | hCoV-19/France/PAC-IHU-0206/2020\|EPI_ISL_569372\|20 | ···················· | ···················· | ····················· | ···················· | ···················· | ···················· |
| K | 703 | hCoV-19/France/PAC-IHU-0208/2020\|EPI_ISL_569373\|20 | ···················· | ···················· | ····················· | ···················· | ···················· | ···················· |
| K | 704 | hCoV-19/France/PAC-IHU-0209/2020\|EPI_ISL_569374\|20 | ···················· | ···················· | ····················· | ···················· | ···················· | ···················· |
| K | 705 | hCoV-19/France/PAC-IHU-0210/2020\|EPI_ISL_569375\|20 | ···················· | ···················· | ····················· | ···················· | ···················· | ···················· |
| K | 706 | hCoV-19/France/PAC-IHU-0212/2020\|EPI_ISL_569377\|20 | ···················· | ···················· | ····················· | ···················· | ···················· | ···················· |
| K | 707 | hCoV-19/France/PAC-IHU-0213/2020\|EPI_ISL_569378\|20 | ···················· | ···················· | ····················· | ···················· | ···················· | ···················· |
| K | 708 | hCoV-19/France/PAC-IHU-0214/2020\|EPI_ISL_569379\|20 | ···················· | ···················· | ····················· | ···················· | ···················· | ···················· |
| K | 709 | hCoV-19/USA/WA-UW-10394/2020\|EPI_ISL_570300\|2020-0 | ···················· | ···················· | ····················· | ···················· | ···················· | ···················· |
| K | 710 | hCoV-19/USA/WA-UW-10396/2020\|EPI_ISL_570301\|2020-0 | ···················· | ···················· | ····················· | ···················· | ···················· | ···················· |
| K | 711 | hCoV-19/USA/WA-UW-10402/2020\|EPI_ISL_570302\|2020-0 | ···················· | ···················· | ····················· | ···················· | ···················· | ···················· |
| K | 712 | hCoV-19/USA/WA-UW-10403/2020\|EPI_ISL_570303\|2020-0 | ···················· | ···················· | ····················· | ···················· | ···················· | ···················· |
| K | 713 | hCoV-19/USA/WA-UW-10408/2020\|EPI_ISL_570304\|2020-0 | ···················· | ···················· | ····················· | ···················· | ···················· | ···················· |
| K | 714 | hCoV-19/USA/WA-UW-10435/2020\|EPI_ISL_570306\|2020-0 | ···················· | ···················· | ····················· | ···················· | ···················· | ···················· |
| K | 715 | hCoV-19/USA/WA-UW-10438/2020\|EPI_ISL_570307\|2020-0 | ···················· | ···················· | ····················· | ···················· | ···················· | ···················· |
| K | 716 | hCoV-19/USA/WA-UW-10439/2020\|EPI_ISL_570308\|2020-0 | ···················· | ···················· | ····················· | ···················· | ···················· | ···················· |
| K | 717 | hCoV-19/USA/WA-UW-10442/2020\|EPI_ISL_570309\|2020-0 | ···················· | ···················· | ····················· | ···················· | ···················· | ···················· |
| K | 718 | hCoV-19/USA/WA-UW-10447/2020\|EPI_ISL_570310\|2020-0 | ···················· | ···················· | ····················· | ···················· | ···················· | ···················· |
| K | 719 | hCoV-19/USA/WA-UW-10449/2020\|EPI_ISL_570311\|2020-0 | ···················· | ···················· | ····················· | ···················· | ···················· | ···················· |
| K | 720 | hCoV-19/USA/WA-UW-10453/2020\|EPI_ISL_570312\|2020-0 | ···················· | ···················· | ····················· | ···················· | ···················· | ···················· |
| K | 721 | hCoV-19/USA/WA-UW-10455/2020\|EPI_ISL_570313\|2020-0 | ···················· | ···················· | ····················· | ···················· | ···················· | ···················· |
| K | 722 | hCoV-19/USA/WA-UW-10459/2020\|EPI_ISL_570314\|2020-0 | ···················· | ···················· | ····················· | ···················· | ···················· | ···················· |
| K | 723 | hCoV-19/USA/WA-UW-10464/2020\|EPI_ISL_570315\|2020-0 | ···················· | ···················· | ····················· | ···················· | ···················· | ···················· |
| K | 724 | hCoV-19/USA/WA-UW-10465/2020\|EPI_ISL_570316\|2020-0 | ···················· | ···················· | ····················· | ···················· | ···················· | ···················· |
| K | 725 | hCoV-19/USA/WA-UW-10471/2020\|EPI_ISL_570317\|2020-0 | ···················· | ···················· | ····················· | ···················· | ···················· | ···················· |
| K | 726 | hCoV-19/USA/WA-UW-10472/2020\|EPI_ISL_570318\|2020-0 | ···················· | ···················· | ····················· | ···················· | ···················· | ···················· |
| K | 727 | hCoV-19/USA/WA-UW-10473/2020\|EPI_ISL_570319\|2020-0 | ···················· | ···················· | ····················· | ···················· | ···················· | ···················· |
| K | 728 | hCoV-19/USA/WA-UW-10479/2020\|EPI_ISL_570320\|2020-0 | ···················· | ···················· | ····················· | ···················· | ···················· | ···················· |
| K | 729 | hCoV-19/USA/WA-UW-10485/2020\|EPI_ISL_570321\|2020-0 | ···················· | ···················· | ····················· | ···················· | ···················· | ···················· |
| K | 730 | hCoV-19/USA/WA-UW-10496/2020\|EPI_ISL_570322\|2020-0 | ···················· | ···················· | ····················· | ···················· | ···················· | ···················· |
| K | 731 | hCoV-19/USA/WA-UW-10501/2020\|EPI_ISL_570323\|2020-0 | ···················· | ···················· | ····················· | ···················· | ···················· | ···················· |
| K | 732 | hCoV-19/USA/WA-UW-10508/2020\|EPI_ISL_570324\|2020-0 | ···················· | ···················· | ····················· | ···················· | ···················· | ···················· |
| K | 733 | hCoV-19/USA/WA-UW-10510/2020\|EPI_ISL_570325\|2020-0 | ···················· | ···················· | ····················· | ···················· | ···················· | ···················· |
| K | 734 | hCoV-19/USA/WA-UW-10511/2020\|EPI_ISL_570326\|2020-0 | ···················· | ···················· | ····················· | ···················· | ···················· | ···················· |
| K | 735 | hCoV-19/USA/WA-UW-10512/2020\|EPI_ISL_570327\|2020-0 | ···················· | ···················· | ····················· | ···················· | ···················· | ···················· |
| K | 736 | hCoV-19/USA/WA-UW-10521/2020\|EPI_ISL_570328\|2020-0 | ···················· | ···················· | ····················· | ···················· | ···················· | ···················· |
| K | 737 | hCoV-19/USA/WA-UW-10523/2020\|EPI_ISL_570329\|2020-0 | ···················· | ···················· | ····················· | ···················· | ···················· | ···················· |
| K | 738 | hCoV-19/USA/WA-UW-10530/2020\|EPI_ISL_570330\|2020-0 | ···················· | ···················· | ····················· | ···················· | ···················· | ···················· |
| K | 739 | hCoV-19/USA/WA-UW-10531/2020\|EPI_ISL_570331\|2020-0 | ···················· | ···················· | ····················· | ···················· | ···················· | ···················· |
| K | 740 | hCoV-19/USA/WA-UW-10533/2020\|EPI_ISL_570332\|2020-0 | ···················· | ···················· | ····················· | ···················· | ···················· | ···················· |
| K | 741 | hCoV-19/USA/WA-UW-10552/2020\|EPI_ISL_570333\|2020-0 | ···················· | ···················· | ····················· | ···················· | ···················· | ···················· |
| K | 742 | hCoV-19/USA/WA-UW-10588/2020\|EPI_ISL_570334\|2020-0 | ···················· | ···················· | ····················· | ···················· | ···················· | ···················· |
| K | 743 | hCoV-19/USA/WA-UW-10599/2020\|EPI_ISL_570335\|2020-0 | ···················· | ···················· | ····················· | ···················· | ···················· | ···················· |
| K | 744 | hCoV-19/USA/WA-UW-10609/2020\|EPI_ISL_570336\|2020-0 | ···················· | ···················· | ····················· | ···················· | ···················· | ···················· |
| K | 745 | hCoV-19/USA/WA-UW-10612/2020\|EPI_ISL_570337\|2020-0 | ···················· | ·A·················· | ····················· | ···················· | ···················· | ···················· |
| K | 746 | hCoV-19/USA/WA-UW-10621/2020\|EPI_ISL_570338\|2020-0 | ···················· | ···················· | ····················· | ···················· | ···················· | ···················· |
| K | 747 | hCoV-19/USA/WA-UW-10648/2020\|EPI_ISL_570339\|2020-0 | ···················· | ···················· | ····················· | ···················· | ···················· | ···················· |
| K | 748 | hCoV-19/USA/WA-UW-10652/2020\|EPI_ISL_570340\|2020-0 | ···················· | ···················· | ····················· | ···················· | ···················· | ···················· |
| K | 749 | hCoV-19/USA/WA-UW-10653/2020\|EPI_ISL_570341\|2020-0 | ···················· | ···················· | ····················· | ···················· | ···················· | ···················· |
| K | 750 | hCoV-19/USA/WA-UW-10654/2020\|EPI_ISL_570342\|2020-0 | ···················· | ···················· | ····················· | ···················· | ···················· | ···················· |
| K | 751 | hCoV-19/USA/WA-UW-10655/2020\|EPI_ISL_570343\|2020-0 | ···················· | ···················· | ····················· | ···················· | ···················· | ···················· |
| K | 752 | hCoV-19/USA/WA-UW-10658/2020\|EPI_ISL_570344\|2020-0 | ···················· | ···················· | ····················· | ···················· | ···················· | ···················· |
| K | 753 | hCoV-19/USA/WA-UW-10663/2020\|EPI_ISL_570345\|2020-0 | ···················· | ···················· | ····················· | ···················· | ···················· | ···················· |
| K | 754 | hCoV-19/USA/WA-UW-10669/2020\|EPI_ISL_570346\|2020-0 | ········T··········· | ·A·················· | ····················· | ···················· | ···················· | ···················· |
| K | 755 | hCoV-19/USA/WA-UW-10685/2020\|EPI_ISL_570348\|2020-0 | ···················· | ···················· | ····················· | ···················· | ···················· | ···················· |
| K | 756 | hCoV-19/USA/WA-UW-10687/2020\|EPI_ISL_570349\|2020-0 | ···················· | ···················· | ····················· | ···················· | ···················· | ···················· |
| K | 757 | hCoV-19/USA/WA-UW-10688/2020\|EPI_ISL_570350\|2020-0 | ···················· | ···················· | ····················· | ···················· | ···················· | ···················· |
| K | 758 | hCoV-19/USA/WA-UW-10691/2020\|EPI_ISL_570351\|2020-0 | ···················· | ···················· | ····················· | ···················· | ···················· | ···················· |
| K | 759 | hCoV-19/USA/WA-UW-10696/2020\|EPI_ISL_570352\|2020-0 | ···················· | ···················· | ····················· | ···················· | ···················· | ···················· |
| K | 760 | hCoV-19/USA/WA-UW-10697/2020\|EPI_ISL_570353\|2020-0 | ···················· | ···················· | ····················· | ···················· | ···················· | ···················· |
| K | 761 | hCoV-19/USA/WA-UW-10698/2020\|EPI_ISL_570354\|2020-0 | ···················· | ···················· | ····················· | ···················· | ···················· | ···················· |
| K | 762 | hCoV-19/USA/WA-UW-10766/2020\|EPI_ISL_570355\|2020-0 | ···················· | ···················· | ····················· | ···················· | ···················· | ···················· |
| K | 763 | hCoV-19/USA/WA-UW-10769/2020\|EPI_ISL_570356\|2020-0 | ···················· | ···················· | ····················· | ···················· | ···················· | ···················· |
| K | 764 | hCoV-19/USA/WA-UW-10818/2020\|EPI_ISL_570358\|2020-0 | ···················· | ···················· | ····················· | ···················· | ···················· | ···················· |
| K | 765 | hCoV-19/USA/WA-UW-10826/2020\|EPI_ISL_570359\|2020-0 | ···················· | ···················· | ····················· | ···················· | ···················· | ···················· |
| K | 766 | hCoV-19/USA/WA-UW-10831/2020\|EPI_ISL_570360\|2020-0 | ···················· | ···················· | ····················· | ···················· | ···················· | ···················· |
| K | 767 | hCoV-19/USA/WA-UW-10834/2020\|EPI_ISL_570361\|2020-0 | ···················· | ···················· | ····················· | ···················· | ···················· | ···················· |

| | | | | | | | | |
|---|---|---|---|---|---|---|---|---|
| K | 768 hCoV-19/USA/WA-UW-10953/2020\|EPI_ISL_570362\|2020-0 | ···················· | ···················· | ···················· | ···················· | ···················· | ···················· | |
| K | 769 hCoV-19/USA/UT-UPHL-03057/2020\|EPI_ISL_560903\|2020 | ···················· | ···················· | ···················· | ···················· | ···················· | ···················· | |
| K | 770 hCoV-19/USA/UT-UPHL-03068/2020\|EPI_ISL_560907\|2020 | ···················· | ···················· | ···················· | ···················· | ···················· | ···················· | |
| K | 771 hCoV-19/USA/UT-UPHL-03071/2020\|EPI_ISL_560908\|2020 | ···················· | ···················· | ···················· | ···················· | ···················· | ···················· | |
| K | 772 hCoV-19/USA/UT-UPHL-03074/2020\|EPI_ISL_560912\|2020 | ···················· | ···················· | ···················· | ···················· | ···················· | ···················· | |
| K | 773 hCoV-19/USA/UT-UPHL-03078/2020\|EPI_ISL_560913\|2020 | ···················· | ···················· | ···················· | ···················· | ···················· | ···················· | |
| K | 774 hCoV-19/USA/UT-UPHL-03076/2020\|EPI_ISL_560914\|2020 | ···················· | ···················· | ···················· | ···················· | ···················· | ···················· | |
| K | 775 hCoV-19/USA/UT-UPHL-03079/2020\|EPI_ISL_560916\|2020 | ···················· | ···················· | ···················· | ···················· | ···················· | ···················· | |
| K | 776 hCoV-19/USA/UT-UPHL-03080/2020\|EPI_ISL_560917\|2020 | ···················· | ···················· | ···················· | ···················· | ···················· | ···················· | |
| K | 777 hCoV-19/USA/UT-UPHL-03087/2020\|EPI_ISL_560918\|2020 | ···················· | ···················· | ···················· | ···················· | ···················· | ···················· | |
| K | 778 hCoV-19/USA/UT-UPHL-03789/2020\|EPI_ISL_560920\|2020 | ···················· | ···················· | ···················· | ···················· | ···················· | ···················· | |
| K | 779 hCoV-19/USA/UT-UPHL-03791/2020\|EPI_ISL_560921\|2020 | ···················· | ···················· | ···················· | ···················· | ···················· | ···················· | |
| K | 780 hCoV-19/USA/UT-UPHL-03793/2020\|EPI_ISL_560922\|2020 | ···················· | ···················· | ···················· | ···················· | ···················· | ···················· | |
| K | 781 hCoV-19/USA/UT-UPHL-03795/2020\|EPI_ISL_560923\|2020 | ···················· | ···················· | ···················· | ···················· | ···················· | ···················· | |
| K | 782 hCoV-19/USA/UT-UPHL-03797/2020\|EPI_ISL_560924\|2020 | ···················· | ···················· | ···················· | ···················· | ···················· | ···················· | |
| K | 783 hCoV-19/USA/TX-DSHS-0481/2020\|EPI_ISL_560926\|2020- | ···················· | ···················· | ···················· | ···················· | ···················· | ···················· | |
| K | 784 hCoV-19/USA/TX-DSHS-0483/2020\|EPI_ISL_560927\|2020- | ···················· | ···················· | ···················· | ···················· | ···················· | ···················· | |
| K | 785 hCoV-19/USA/TX-DSHS-0485/2020\|EPI_ISL_560929\|2020- | ···················· | ···················· | ···················· | ···················· | ···················· | ···················· | |
| K | 786 hCoV-19/USA/TX-DSHS-0486/2020\|EPI_ISL_560930\|2020- | ···················· | ···················· | ···················· | ···················· | ···················· | ···················· | |
| K | 787 hCoV-19/USA/TX-DSHS-0487/2020\|EPI_ISL_560931\|2020- | ···················· | ···················· | ···················· | ···················· | ···················· | ···················· | |
| K | 788 hCoV-19/USA/TX-DSHS-0488/2020\|EPI_ISL_560932\|2020- | ···················· | ···················· | ···················· | ···················· | ···················· | ···················· | |
| K | 789 hCoV-19/USA/TX-DSHS-0489/2020\|EPI_ISL_560933\|2020- | ···················· | ···················· | ···················· | ···················· | ···················· | ···················· | |
| K | 790 hCoV-19/USA/TX-DSHS-0491/2020\|EPI_ISL_560935\|2020- | ···················· | ···················· | ···················· | ···················· | ···················· | ···················· | |
| K | 791 hCoV-19/USA/TX-DSHS-0492/2020\|EPI_ISL_560936\|2020- | ···················· | ·A·················· | ···················· | ···················· | ···················· | ···················· | |
| K | 792 hCoV-19/USA/TX-DSHS-0493/2020\|EPI_ISL_560937\|2020- | ···················· | ···················· | ···················· | ···················· | ···················· | ···················· | |
| K | 793 hCoV-19/USA/TX-DSHS-0494/2020\|EPI_ISL_560938\|2020- | ···················· | ···················· | ···················· | ···················· | ···················· | ···················· | |
| K | 794 hCoV-19/USA/TX-DSHS-0495/2020\|EPI_ISL_560939\|2020- | ···················· | ···················· | ···················· | ···················· | ···················· | ···················· | |
| K | 795 hCoV-19/USA/TX-DSHS-0497/2020\|EPI_ISL_560941\|2020- | ···················· | ···················· | ···················· | ···················· | ···················· | ···················· | |
| K | 796 hCoV-19/USA/TX-DSHS-0500/2020\|EPI_ISL_560944\|2020- | ···················· | ·A·················· | ···················· | ···················· | ···················· | ···················· | |
| K | 797 hCoV-19/USA/TX-DSHS-0503/2020\|EPI_ISL_560947\|2020- | ···················· | ···················· | ···················· | ···················· | ···················· | ···················· | |
| K | 798 hCoV-19/USA/TX-DSHS-0504/2020\|EPI_ISL_560948\|2020- | ···················· | ·A·················· | ···················· | ···················· | ···················· | ···················· | |
| K | 799 hCoV-19/USA/TX-DSHS-0505/2020\|EPI_ISL_560949\|2020- | ···················· | ···················· | ···················· | ···················· | ···················· | ···················· | |
| K | 800 hCoV-19/USA/TX-DSHS-0508/2020\|EPI_ISL_560951\|2020- | ···················· | ·A·················· | ···················· | ···················· | ···················· | ···················· | |
| K | 801 hCoV-19/USA/TX-DSHS-0512/2020\|EPI_ISL_560955\|2020- | ···················· | ···················· | ···················· | ···················· | ···················· | ···················· | |
| K | 802 hCoV-19/USA/TX-DSHS-0516/2020\|EPI_ISL_560959\|2020- | ···················· | ···················· | ···················· | ···················· | ···················· | ···················· | |
| K | 803 hCoV-19/Australia/VIC10936/2020\|EPI_ISL_563160\|202 | ···················· | ···················· | ···················· | ···················· | ···················· | ···················· | |
| K | 804 hCoV-19/Australia/VIC10981/2020\|EPI_ISL_563180\|202 | ···················· | ···················· | ···················· | ···················· | ···················· | ···················· | |
| K | 805 hCoV-19/Australia/VIC10982/2020\|EPI_ISL_563181\|202 | ···················· | ···················· | ···················· | ···················· | ···················· | ···················· | |
| K | 806 hCoV-19/Australia/VIC10983/2020\|EPI_ISL_563182\|202 | ···················· | ···················· | ···················· | ···················· | ···················· | ···················· | |
| K | 807 hCoV-19/Australia/VIC10985/2020\|EPI_ISL_563183\|202 | ···················· | ···················· | ···················· | ···················· | ···················· | ···················· | |
| K | 808 hCoV-19/Australia/VIC10986/2020\|EPI_ISL_563184\|202 | ···················· | ···················· | ···················· | ···················· | ···················· | ···················· | |
| K | 809 hCoV-19/Australia/VIC10989/2020\|EPI_ISL_563185\|202 | ···················· | ···················· | ···················· | ···················· | ···················· | ···················· | |
| K | 810 hCoV-19/Australia/VIC10991/2020\|EPI_ISL_563186\|202 | ···················· | ···················· | ···················· | ···················· | ···················· | ···················· | |
| K | 811 hCoV-19/Australia/VIC10992/2020\|EPI_ISL_563187\|202 | ···················· | ···················· | ···················· | ···················· | ···················· | ···················· | |
| K | 812 hCoV-19/Australia/VIC10993/2020\|EPI_ISL_563188\|202 | ···················· | ···················· | ···················· | ···················· | ···················· | ···················· | |
| K | 813 hCoV-19/Australia/VIC10996/2020\|EPI_ISL_563189\|202 | ···················· | ···················· | ···················· | ···················· | NNN················· | ···················· | 1 |
| K | 815 hCoV-19/Australia/VIC10999/2020\|EPI_ISL_563191\|202 | ···················· | ···················· | ···················· | ···················· | ···················· | ···················· | |
| K | 816 hCoV-19/Australia/VIC11000/2020\|EPI_ISL_563192\|202 | ···················· | ···················· | ···················· | ···················· | ···················· | ···················· | |
| K | 817 hCoV-19/Australia/VIC11004/2020\|EPI_ISL_563193\|202 | ···················· | ···················· | ···················· | ···················· | ···················· | ···················· | |
| K | 819 hCoV-19/Australia/VIC11007/2020\|EPI_ISL_563195\|202 | ···················· | ···················· | ···················· | ···················· | ···················· | ···················· | |
| K | 820 hCoV-19/Australia/VIC11011/2020\|EPI_ISL_563196\|202 | ···················· | ···················· | ···················· | ···················· | ···················· | ···················· | |
| K | 822 hCoV-19/Australia/VIC11013/2020\|EPI_ISL_563198\|202 | ···················· | ···················· | ···················· | ···················· | ···················· | ···················· | |
| K | 823 hCoV-19/Australia/VIC11014/2020\|EPI_ISL_563199\|202 | ···················· | ···················· | ···················· | ···················· | ···················· | ···················· | |
| K | 824 hCoV-19/Australia/VIC11017/2020\|EPI_ISL_563200\|202 | ···················· | ···················· | ···················· | ···················· | ···················· | ···················· | |
| K | 825 hCoV-19/Australia/VIC11018/2020\|EPI_ISL_563201\|202 | ···················· | ···················· | ···················· | ···················· | ···················· | ···················· | |
| K | 826 hCoV-19/Australia/VIC11019/2020\|EPI_ISL_563202\|202 | ···················· | ···················· | ···················· | ···················· | ···················· | ···················· | |
| K | 827 hCoV-19/Australia/VIC11020/2020\|EPI_ISL_563203\|202 | ···················· | ···················· | ···················· | ···················· | ···················· | ···················· | |
| K | 828 hCoV-19/Australia/VIC11021/2020\|EPI_ISL_563204\|202 | ···················· | ···················· | ···················· | ···················· | ···················· | ···················· | |
| K | 830 hCoV-19/Australia/VIC11023/2020\|EPI_ISL_563206\|202 | ···················· | ···················· | ···················· | ···················· | ···················· | ···················· | |
| K | 831 hCoV-19/Australia/VIC11024/2020\|EPI_ISL_563207\|202 | ···················· | ···················· | ···················· | ···················· | ···················· | ···················· | |
| K | 832 hCoV-19/Australia/VIC11025/2020\|EPI_ISL_563208\|202 | ···················· | ···················· | ···················· | ···················· | ···················· | ···················· | |
| K | 833 hCoV-19/Australia/VIC11028/2020\|EPI_ISL_563209\|202 | ···················· | ···················· | ···················· | ···················· | ···················· | ···················· | |
| K | 834 hCoV-19/Australia/VIC11030/2020\|EPI_ISL_563210\|202 | ···················· | ···················· | ···················· | ···················· | ···················· | ···················· | |
| K | 835 hCoV-19/Australia/VIC11032/2020\|EPI_ISL_563211\|202 | ···················· | ···················· | ···················· | ···················· | ···················· | ···················· | |
| K | 836 hCoV-19/Australia/VIC11033/2020\|EPI_ISL_563212\|202 | ···················· | ···················· | ···················· | ···················· | ···················· | ···················· | |
| K | 838 hCoV-19/Australia/VIC11040/2020\|EPI_ISL_563214\|202 | ···················· | ···················· | ···················· | ···················· | ···················· | ···················· | |
| K | 839 hCoV-19/Australia/VIC11041/2020\|EPI_ISL_563215\|202 | ···················· | ···················· | ···················· | ···················· | ···················· | ···················· | |
| K | 840 hCoV-19/Australia/VIC11042/2020\|EPI_ISL_563216\|202 | ···················· | ···················· | ···················· | ···················· | ···················· | ···················· | |
| K | 841 hCoV-19/Australia/VIC11043/2020\|EPI_ISL_563217\|202 | ···················· | ···················· | ···················· | ···················· | ···················· | ···················· | |
| K | 842 hCoV-19/Australia/VIC11044/2020\|EPI_ISL_563218\|202 | ···················· | ···················· | ···················· | ···················· | ···················· | ···················· | |
| K | 843 hCoV-19/Australia/VIC11045/2020\|EPI_ISL_563219\|202 | ···················· | ···················· | ···················· | ···················· | ···················· | ···················· | |
| K | 844 hCoV-19/Australia/VIC11047/2020\|EPI_ISL_563220\|202 | ···················· | ···················· | ···················· | ···················· | ···················· | ···················· | |
| K | 846 hCoV-19/Australia/VIC11050/2020\|EPI_ISL_563222\|202 | ···················· | ···················· | ···················· | ···················· | ···················· | ···················· | |
| K | 847 hCoV-19/Australia/VIC11051/2020\|EPI_ISL_563223\|202 | ···················· | ···················· | ···················· | ···················· | ···················· | ···················· | |
| K | 848 hCoV-19/Australia/VIC11052/2020\|EPI_ISL_563224\|202 | ···················· | ···················· | ···················· | ···················· | ···················· | ···················· | |
| K | 849 hCoV-19/Australia/VIC11054/2020\|EPI_ISL_563225\|202 | ···················· | ···················· | ···················· | ···················· | ···················· | ···················· | |
| K | 850 hCoV-19/Australia/VIC11056/2020\|EPI_ISL_563226\|202 | ···················· | ···················· | ···················· | ···················· | ···················· | ···················· | |
| K | 852 hCoV-19/Australia/VIC11061/2020\|EPI_ISL_563228\|202 | ···················· | ···················· | ···················· | ···················· | ···················· | ···················· | |
| K | 853 hCoV-19/Australia/VIC11067/2020\|EPI_ISL_563229\|202 | ···················· | ···················· | ···················· | ···················· | ···················· | ···················· | |
| K | 854 hCoV-19/Australia/VIC11068/2020\|EPI_ISL_563230\|202 | ···················· | ···················· | ···················· | ···················· | ···················· | ···················· | |

| | | | | | | | | |
|---|---|---|---|---|---|---|---|---|
| K | 855 hCoV-19/Australia/VIC11071/2020\|EPI_ISL_563231\|202 | ···················· | ···················· | ···················· | ···················· | ···················· | ···················· | |
| K | 856 hCoV-19/Australia/VIC11073/2020\|EPI_ISL_563232\|202 | ···················· | ···················· | ···················· | ···················· | ···················· | ···················· | |
| K | 857 hCoV-19/Australia/VIC11074/2020\|EPI_ISL_563233\|202 | ···················· | ···················· | ···················· | ···················· | ···················· | ···················· | |
| K | 858 hCoV-19/Australia/VIC11075/2020\|EPI_ISL_563234\|202 | ···················· | ···················· | ···················· | ···················· | ···················· | ···················· | |
| K | 860 hCoV-19/Australia/VIC11077/2020\|EPI_ISL_563236\|202 | ···················· | ···················· | ···················· | ···················· | ···················· | ···················· | |
| K | 861 hCoV-19/Australia/VIC11081/2020\|EPI_ISL_563237\|202 | ···················· | ···················· | ···················· | ···················· | ···················· | ···················· | |
| K | 863 hCoV-19/Australia/VIC11083/2020\|EPI_ISL_563239\|202 | ···················· | ···················· | ···················· | ···················· | ···················· | ···················· | |
| K | 865 hCoV-19/Australia/VIC11085/2020\|EPI_ISL_563241\|202 | ···················· | ···················· | ···················· | ···················· | ···················· | ···················· | |
| K | 866 hCoV-19/Australia/VIC11086/2020\|EPI_ISL_563242\|202 | ···················· | ···················· | ···················· | ···················· | ···················· | ···················· | |
| K | 867 hCoV-19/Australia/VIC11088/2020\|EPI_ISL_563243\|202 | ···················· | ···················· | ···················· | ···················· | ···················· | ···················· | |
| K | 868 hCoV-19/Australia/VIC11089/2020\|EPI_ISL_563244\|202 | ···················· | ···················· | ···················· | ···················· | ···················· | ···················· | |
| K | 869 hCoV-19/Australia/VIC11099/2020\|EPI_ISL_563245\|202 | ···················· | ···················· | ···················· | ···················· | ···················· | ···················· | |
| K | 870 hCoV-19/Australia/VIC11100/2020\|EPI_ISL_563246\|202 | ···················· | ···················· | ···················· | ···················· | ···················· | ···················· | |
| K | 871 hCoV-19/Australia/VIC11101/2020\|EPI_ISL_563247\|202 | ···················· | ···················· | ···················· | ···················· | ···················· | ···················· | |
| K | 872 hCoV-19/Australia/VIC11102/2020\|EPI_ISL_563248\|202 | ···················· | ···················· | ···················· | ···················· | ···················· | ···················· | |
| K | 873 hCoV-19/Australia/VIC11103/2020\|EPI_ISL_563249\|202 | ···················· | ···················· | ···················· | ···················· | ···················· | ···················· | |
| K | 874 hCoV-19/Australia/VIC11104/2020\|EPI_ISL_563250\|202 | ···················· | ···················· | ···················· | ···················· | ···················· | ···················· | |
| K | 875 hCoV-19/Australia/VIC11106/2020\|EPI_ISL_563251\|202 | ···················· | ···················· | ···················· | ···················· | ···················· | ···················· | |
| K | 876 hCoV-19/Australia/VIC11109/2020\|EPI_ISL_563252\|202 | ···················· | ···················· | ···················· | ···················· | ···················· | ···················· | |
| K | 877 hCoV-19/Australia/VIC11112/2020\|EPI_ISL_563253\|202 | ···················· | ···················· | ···················· | ···················· | ···················· | ···················· | |
| K | 878 hCoV-19/Australia/VIC11114/2020\|EPI_ISL_563254\|202 | ···················· | ···················· | ···················· | ···················· | ···················· | ···················· | |
| K | 879 hCoV-19/Australia/VIC11115/2020\|EPI_ISL_563255\|202 | ···················· | ···················· | ···················· | ···················· | ···················· | ···················· | |
| K | 880 hCoV-19/Australia/VIC11119/2020\|EPI_ISL_563256\|202 | ···················· | ···················· | ···················· | ···················· | ···················· | ···················· | |
| K | 881 hCoV-19/Australia/VIC11120/2020\|EPI_ISL_563257\|202 | ···················· | ···················· | ···················· | ···················· | ···················· | ···················· | |
| K | 882 hCoV-19/Australia/VIC11122/2020\|EPI_ISL_563258\|202 | ···················· | ···················· | ···················· | ···················· | ···················· | ···················· | |
| K | 883 hCoV-19/Australia/VIC11127/2020\|EPI_ISL_563259\|202 | ···················· | ···················· | ···················· | ···················· | ···················· | ···················· | |
| K | 885 hCoV-19/Australia/VIC11130/2020\|EPI_ISL_563261\|202 | ···················· | ···················· | ···················· | ···················· | ···················· | ···················· | |
| K | 886 hCoV-19/Australia/VIC11132/2020\|EPI_ISL_563262\|202 | ···················· | ···················· | ···················· | ···················· | ···················· | ···················· | |
| K | 887 hCoV-19/Australia/VIC11134/2020\|EPI_ISL_563263\|202 | ···················· | ···················· | ···················· | ···················· | ···················· | ···················· | |
| K | 888 hCoV-19/Australia/VIC11135/2020\|EPI_ISL_563264\|202 | ···················· | ···················· | ···················· | ···················· | ···················· | ···················· | |
| K | 889 hCoV-19/Australia/VIC11136/2020\|EPI_ISL_563265\|202 | ···················· | ···················· | ···················· | ···················· | ···················· | ···················· | |
| K | 890 hCoV-19/Australia/VIC11140/2020\|EPI_ISL_563266\|202 | ···················· | ···················· | ···················· | ···················· | ···················· | ···················· | |
| K | 891 hCoV-19/Australia/VIC11141/2020\|EPI_ISL_563267\|202 | ···················· | ···················· | ···················· | ···················· | ···················· | ···················· | |
| K | 892 hCoV-19/Australia/VIC11143/2020\|EPI_ISL_563268\|202 | ···················· | ···················· | ···················· | ···················· | ···················· | ···················· | |
| K | 893 hCoV-19/Australia/VIC11146/2020\|EPI_ISL_563269\|202 | ···················· | ···················· | ···················· | ···················· | ···················· | ···················· | |
| K | 894 hCoV-19/Australia/VIC11147/2020\|EPI_ISL_563270\|202 | ···················· | ···················· | ···················· | ···················· | ···················· | ···················· | |
| K | 895 hCoV-19/Australia/VIC11148/2020\|EPI_ISL_563271\|202 | ···················· | ···················· | ···················· | ···················· | ···················· | ···················· | |
| K | 896 hCoV-19/Australia/VIC11152/2020\|EPI_ISL_563272\|202 | ···················· | ···················· | ···················· | ···················· | ···················· | ···················· | |
| K | 897 hCoV-19/Australia/VIC11153/2020\|EPI_ISL_563273\|202 | ···················· | ·····R·············· | ···················· | ···················· | ···················· | ···················· | |
| K | 898 hCoV-19/Australia/VIC11155/2020\|EPI_ISL_563274\|202 | ···················· | ···················· | ···················· | ···················· | ···················· | ···················· | |
| K | 899 hCoV-19/Australia/VIC11156/2020\|EPI_ISL_563275\|202 | ···················· | ···················· | ···················· | ···················· | ···················· | ···················· | |
| K | 900 hCoV-19/Australia/VIC11158/2020\|EPI_ISL_563276\|202 | ···················· | ···················· | ···················· | ···················· | ···················· | ···················· | |
| K | 901 hCoV-19/Australia/VIC11162/2020\|EPI_ISL_563277\|202 | ···················· | ···················· | ···················· | ···················· | ···················· | ···················· | |
| K | 902 hCoV-19/Australia/VIC11163/2020\|EPI_ISL_563278\|202 | ···················· | ···················· | ···················· | ···················· | ···················· | ···················· | |
| K | 903 hCoV-19/Australia/VIC7548/2020\|EPI_ISL_564491\|2020 | ···················· | ···················· | ···················· | ···················· | ···················· | ···················· | |
| K | 904 hCoV-19/Australia/VIC7598/2020\|EPI_ISL_564522\|2020 | ···················· | ···················· | ···················· | ···················· | ···················· | ···················· | |
| K | 905 hCoV-19/Australia/VIC7603/2020\|EPI_ISL_564523\|2020 | ···················· | ···················· | ···················· | ···················· | ···················· | ···················· | |
| K | 906 hCoV-19/Australia/VIC7604/2020\|EPI_ISL_564524\|2020 | ···················· | ···················· | ···················· | ···················· | ···················· | ···················· | |
| K | 907 hCoV-19/Australia/VIC7609/2020\|EPI_ISL_564525\|2020 | ···················· | ···················· | ···················· | ···················· | ···················· | ···················· | |
| K | 908 hCoV-19/Australia/VIC7610/2020\|EPI_ISL_564526\|2020 | ···················· | ···················· | ···················· | ···················· | ···················· | ···················· | |
| K | 909 hCoV-19/Australia/VIC7611/2020\|EPI_ISL_564527\|2020 | ···················· | ···················· | ···················· | ···················· | ···················· | ···················· | |
| K | 910 hCoV-19/Australia/VIC7612/2020\|EPI_ISL_564528\|2020 | ···················· | ···················· | ···················· | ···················· | ···················· | ···················· | |
| K | 911 hCoV-19/Australia/VIC7613/2020\|EPI_ISL_564529\|2020 | ···················· | ···················· | ···················· | ···················· | ···················· | ···················· | |
| K | 912 hCoV-19/Australia/VIC7614/2020\|EPI_ISL_564530\|2020 | ···················· | ···················· | ···················· | ···················· | ···················· | ···················· | |
| K | 913 hCoV-19/Australia/VIC7615/2020\|EPI_ISL_564531\|2020 | ···················· | ···················· | ···················· | ···················· | ···················· | ···················· | |
| K | 914 hCoV-19/Australia/VIC7616/2020\|EPI_ISL_564532\|2020 | ···················· | ···················· | ···················· | ···················· | ···················· | ···················· | |
| K | 915 hCoV-19/Australia/VIC7617/2020\|EPI_ISL_564533\|2020 | ···················· | ···················· | ···················· | ···················· | ···················· | ···················· | |
| K | 916 hCoV-19/Australia/VIC7618/2020\|EPI_ISL_564534\|2020 | ···················· | ···················· | ···················· | ···················· | ···················· | ···················· | |
| K | 917 hCoV-19/Australia/VIC7619/2020\|EPI_ISL_564535\|2020 | ···················· | ···················· | ···················· | ···················· | ···················· | ···················· | |
| K | 918 hCoV-19/Australia/VIC7620/2020\|EPI_ISL_564536\|2020 | ···················· | ···················· | ···················· | ···················· | ···················· | ···················· | |
| K | 919 hCoV-19/Australia/VIC7621/2020\|EPI_ISL_564537\|2020 | ···················· | ···················· | ···················· | ···················· | ···················· | ···················· | |
| K | 920 hCoV-19/Australia/VIC7622/2020\|EPI_ISL_564538\|2020 | ···················· | ···················· | ···················· | ···················· | ···················· | ···················· | |
| K | 921 hCoV-19/Australia/VIC7623/2020\|EPI_ISL_564539\|2020 | ···················· | ···················· | ···················· | ···················· | ···················· | ···················· | |
| K | 922 hCoV-19/Australia/VIC7624/2020\|EPI_ISL_564540\|2020 | ···················· | ···················· | ···················· | ···················· | ···················· | ···················· | |
| K | 923 hCoV-19/Australia/VIC7625/2020\|EPI_ISL_564541\|2020 | ···················· | ···················· | ···················· | ···················· | ···················· | ···················· | |
| K | 924 hCoV-19/Australia/VIC7627/2020\|EPI_ISL_564542\|2020 | ···················· | ···················· | ···················· | ···················· | ···················· | ···················· | |
| K | 925 hCoV-19/Australia/VIC7628/2020\|EPI_ISL_564543\|2020 | ···················· | ···················· | ···················· | ···················· | ···················· | ·················M·· | 1 |
| K | 926 hCoV-19/Australia/VIC7639/2020\|EPI_ISL_564544\|2020 | ···················· | ···················· | ···················· | ···················· | ···················· | ···················· | |
| K | 927 hCoV-19/Australia/VIC7640/2020\|EPI_ISL_564545\|2020 | ···················· | ···················· | ···················· | ···················· | ···················· | ···················· | |
| K | 928 hCoV-19/Australia/VIC7641/2020\|EPI_ISL_564546\|2020 | ···················· | ···················· | ···················· | ···················· | ···················· | ···················· | |
| K | 929 hCoV-19/Australia/VIC7643/2020\|EPI_ISL_564547\|2020 | ···················· | ···················· | ···················· | ···················· | ···················· | ···················· | |
| K | 930 hCoV-19/Australia/VIC7644/2020\|EPI_ISL_564548\|2020 | ···················· | ···················· | ··K················· | ···················· | ···················· | ···················· | |
| K | 931 hCoV-19/Australia/VIC7646/2020\|EPI_ISL_564549\|2020 | ···················· | ···················· | ··K················· | ···················· | ···················· | ···················· | |
| K | 932 hCoV-19/Australia/VIC7648/2020\|EPI_ISL_564550\|2020 | ···················· | ···················· | ··K················· | ···················· | ···················· | ···················· | |
| K | 933 hCoV-19/Australia/VIC7650/2020\|EPI_ISL_564551\|2020 | ···················· | ···················· | ···················· | ···················· | ···················· | ···················· | |
| K | 934 hCoV-19/Australia/VIC7651/2020\|EPI_ISL_564552\|2020 | ···················· | ···················· | ···················· | ···················· | ···················· | ···················· | |
| K | 935 hCoV-19/Australia/VIC7653/2020\|EPI_ISL_564553\|2020 | ···················· | ···················· | ···················· | ···················· | ···················· | ··N················· | |
| K | 936 hCoV-19/Australia/VIC7654/2020\|EPI_ISL_564554\|2020 | ···················· | ···················· | ··K················· | ···················· | ···················· | ···················· | |
| K | 937 hCoV-19/Australia/VIC7655/2020\|EPI_ISL_564555\|2020 | ···················· | ···················· | ··K················· | ···················· | ···················· | ···················· | |
| K | 938 hCoV-19/Australia/VIC7656/2020\|EPI_ISL_564556\|2020 | ···················· | ···················· | ···················· | ···················· | ···················· | ···················· | |

| | | | | | | | | |
|---|---|---|---|---|---|---|---|---|
| K | 939 | hCoV-19/Australia/VIC7657/2020\|EPI_ISL_564557\|2020 | ···················· | ···················· | ···················· | ···················· | ···················· | ···················· |
| K | 940 | hCoV-19/Australia/VIC7658/2020\|EPI_ISL_564558\|2020 | ···················· | ···················· | ···················· | ···················· | ···················· | ···················· |
| K | 941 | hCoV-19/Australia/VIC7659/2020\|EPI_ISL_564559\|2020 | ···················· | ···················· | ···················· | ···················· | ···················· | ···················· |
| K | 942 | hCoV-19/Australia/VIC7660/2020\|EPI_ISL_564560\|2020 | ···················· | ···················· | ···················· | ···················· | ···················· | ···················· |
| K | 943 | hCoV-19/Australia/VIC7662/2020\|EPI_ISL_564561\|2020 | ···················· | ···················· | ···················· | ···················· | ···················· | ···················· |
| K | 944 | hCoV-19/Australia/VIC7665/2020\|EPI_ISL_564562\|2020 | ···················· | ···················· | ···················· | ···················· | ···················· | ···················· |
| K | 946 | hCoV-19/Australia/VIC7667/2020\|EPI_ISL_564564\|2020 | ···················· | ···················· | ···················· | ···················· | ···················· | ···················· |
| K | 947 | hCoV-19/Australia/VIC7670/2020\|EPI_ISL_564565\|2020 | ···················· | ···················· | ···················· | ···················· | ···················· | ···················· |
| K | 948 | hCoV-19/Australia/VIC7671/2020\|EPI_ISL_564566\|2020 | ···················· | ···················· | ···················· | ···················· | ···················· | ···················· |
| K | 949 | hCoV-19/Australia/VIC7672/2020\|EPI_ISL_564567\|2020 | ···················· | ···················· | ···················· | ···················· | ···················· | ···················· |
| K | 950 | hCoV-19/Australia/VIC7675/2020\|EPI_ISL_564568\|2020 | ···················· | ···················· | ···················· | ···················· | ···················· | ···················· |
| K | 951 | hCoV-19/Australia/VIC7676/2020\|EPI_ISL_564569\|2020 | ···················· | ···················· | ···················· | ···················· | ···················· | ···················· |
| K | 952 | hCoV-19/Australia/VIC7677/2020\|EPI_ISL_564570\|2020 | ···················· | ···················· | ···················· | ···················· | ···················· | ···················· |
| K | 953 | hCoV-19/Australia/VIC7678/2020\|EPI_ISL_564571\|2020 | ···················· | ···················· | ···················· | ···················· | ···················· | ···················· |
| K | 954 | hCoV-19/Australia/VIC7681/2020\|EPI_ISL_564572\|2020 | ···················· | ···················· | ···················· | ···················· | ···················· | ···················· |
| K | 955 | hCoV-19/Australia/VIC7683/2020\|EPI_ISL_564573\|2020 | ···················· | ···················· | ···················· | ···················· | ···················· | ···················· |
| K | 956 | hCoV-19/Australia/VIC7684/2020\|EPI_ISL_564574\|2020 | ···················· | ···················· | ···················· | ···················· | ···················· | ···················· |
| K | 957 | hCoV-19/Australia/VIC7685/2020\|EPI_ISL_564575\|2020 | ···················· | ···················· | ···················· | ···················· | ···················· | ···················· |
| K | 958 | hCoV-19/Australia/VIC7686/2020\|EPI_ISL_564576\|2020 | ···················· | ···················· | ···················· | ···················· | ···················· | ···················· |
| K | 959 | hCoV-19/Australia/VIC7687/2020\|EPI_ISL_564577\|2020 | ···················· | ···················· | ···················· | ···················· | ···················· | ···················· |
| K | 960 | hCoV-19/Australia/VIC7688/2020\|EPI_ISL_564578\|2020 | ···················· | ···················· | ···················· | ···················· | ···················· | ···················· |
| K | 961 | hCoV-19/Australia/VIC7690/2020\|EPI_ISL_564579\|2020 | ···················· | ···················· | ···················· | ···················· | ···················· | ···················· |
| K | 962 | hCoV-19/Australia/VIC7691/2020\|EPI_ISL_564580\|2020 | ···················· | ···················· | ···················· | ···················· | ···················· | ···················· |
| K | 963 | hCoV-19/Australia/VIC7692/2020\|EPI_ISL_564581\|2020 | ···················· | ···················· | ···················· | ···················· | ···················· | ···················· |
| K | 964 | hCoV-19/Australia/VIC7693/2020\|EPI_ISL_564582\|2020 | ···················· | ···················· | ···················· | ···················· | ···················· | ···················· |
| K | 965 | hCoV-19/Australia/VIC7694/2020\|EPI_ISL_564583\|2020 | ···················· | ···················· | ···················· | ···················· | ···················· | ···················· |
| K | 966 | hCoV-19/Australia/VIC7695/2020\|EPI_ISL_564584\|2020 | ···················· | ···················· | ···················· | ···················· | ···················· | ···················· |
| K | 967 | hCoV-19/Australia/VIC7696/2020\|EPI_ISL_564585\|2020 | ···················· | ···················· | ···················· | ···················· | ···················· | ···················· |
| K | 968 | hCoV-19/Australia/VIC7697/2020\|EPI_ISL_564586\|2020 | ···················· | ···················· | ···················· | ···················· | ···················· | ···················· |
| K | 969 | hCoV-19/Australia/VIC7698/2020\|EPI_ISL_564587\|2020 | ···················· | ···················· | ···················· | ···················· | ···················· | ···················· |
| K | 970 | hCoV-19/Australia/VIC7699/2020\|EPI_ISL_564588\|2020 | ···················· | ···················· | ···················· | ···················· | ···················· | ···················· |
| K | 971 | hCoV-19/Australia/VIC7700/2020\|EPI_ISL_564589\|2020 | ···················· | ···················· | ···················· | ···················· | ···················· | ···················· |
| K | 972 | hCoV-19/Australia/VIC7701/2020\|EPI_ISL_564590\|2020 | ···················· | ···················· | ···················· | ···················· | ···················· | ···················· |
| K | 973 | hCoV-19/Australia/VIC7702/2020\|EPI_ISL_564591\|2020 | ···················· | ···················· | ···················· | ···················· | ···················· | ···················· |
| K | 974 | hCoV-19/Australia/VIC7704/2020\|EPI_ISL_564592\|2020 | ···················· | ···················· | ···················· | ···················· | ···················· | ···················· |
| K | 975 | hCoV-19/Australia/VIC7705/2020\|EPI_ISL_564593\|2020 | ···················· | ···················· | ···················· | ···················· | ···················· | ···················· |
| K | 976 | hCoV-19/Australia/VIC7706/2020\|EPI_ISL_564594\|2020 | ···················· | ···················· | ···················· | ···················· | ···················· | ···················· |
| K | 977 | hCoV-19/Australia/VIC7710/2020\|EPI_ISL_564595\|2020 | ···················· | ···················· | ···················· | ···················· | ···················· | ···················· |
| K | 978 | hCoV-19/Australia/VIC7711/2020\|EPI_ISL_564596\|2020 | ···················· | ···················· | ···················· | ···················· | ···················· | ···················· |
| K | 979 | hCoV-19/Australia/VIC7712/2020\|EPI_ISL_564597\|2020 | ···················· | ···················· | ···················· | ···················· | ···················· | ···················· |
| K | 980 | hCoV-19/Australia/VIC7713/2020\|EPI_ISL_564598\|2020 | ···················· | ···················· | ···················· | ···················· | ···················· | ···················· |
| K | 981 | hCoV-19/Australia/VIC7716/2020\|EPI_ISL_564599\|2020 | ···················· | ···················· | ···················· | ···················· | ···················· | ···················· |
| K | 982 | hCoV-19/USA/MI-MDHHS-SC21810/2020\|EPI_ISL_565853\|2 | ···················· | ···················· | ···················· | ···················· | ···················· | ···················· |
| K | 984 | hCoV-19/USA/MI-MDHHS-SC21830/2020\|EPI_ISL_565873\|2 | ···················· | ···················· | ···················· | ···················· | ···················· | ···················· |
| K | 985 | hCoV-19/USA/MI-MDHHS-SC21838/2020\|EPI_ISL_565881\|2 | ···················· | ···················· | ···················· | ···················· | ···················· | ···················· |
| K | 986 | hCoV-19/USA/MI-MDHHS-SC21841/2020\|EPI_ISL_565884\|2 | ···················· | ···················· | ···················· | ···················· | ···················· | ···················· |
| K | 987 | hCoV-19/England/ALDP-9BE9A3/2020\|EPI_ISL_568056\|20 | ···················· | ···················· | ···················· | ···················· | ···················· | ···················· |
| K | 988 | hCoV-19/England/ALDP-9BE3D2/2020\|EPI_ISL_568057\|20 | ···················· | ···················· | ···················· | ···················· | ···················· | ···················· |
| K | 989 | hCoV-19/England/ALDP-9BEC8C/2020\|EPI_ISL_568058\|20 | ···················· | ···················· | ···················· | ···················· | ···················· | ···················· |
| K | 990 | hCoV-19/England/ALDP-9BDF75/2020\|EPI_ISL_568059\|20 | ···················· | ···················· | ···················· | ···················· | ···················· | ···················· |
| K | 991 | hCoV-19/England/ALDP-9BE581/2020\|EPI_ISL_568060\|20 | ···················· | ···················· | ···················· | ···················· | ···················· | ···················· |
| K | 992 | hCoV-19/England/ALDP-9BDDB7/2020\|EPI_ISL_568061\|20 | ···················· | ···················· | ···················· | ···················· | ···················· | ···················· |
| K | 993 | hCoV-19/England/ALDP-9BE7C7/2020\|EPI_ISL_568063\|20 | ···················· | ···················· | ···················· | ···················· | ···················· | ···················· |
| K | 994 | hCoV-19/England/ALDP-9BDED2/2020\|EPI_ISL_568064\|20 | ···················· | ···················· | ···················· | ···················· | ···················· | ···················· |
| K | 995 | hCoV-19/England/ALDP-9BC987/2020\|EPI_ISL_568065\|20 | ···················· | ···················· | ···················· | ···················· | ···················· | ···················· |
| K | 996 | hCoV-19/England/ALDP-9BE080/2020\|EPI_ISL_568066\|20 | ···················· | ···················· | ···················· | ···················· | ···················· | ···················· |
| K | 997 | hCoV-19/England/ALDP-9BC732/2020\|EPI_ISL_568067\|20 | ···················· | ···················· | ···················· | ···················· | ···················· | ···················· |
| K | 998 | hCoV-19/England/ALDP-9BDD02/2020\|EPI_ISL_568069\|20 | ···················· | ·A·················· | ···················· | ···················· | ···················· | ··A················· |
| K | 999 | hCoV-19/France/PAC-IHU-0224/2020\|EPI_ISL_569387\|20 | ···················· | ···················· | ···················· | ···················· | ···················· | ···················· |
| K | 1000 | hCoV-19/France/PAC-IHU-0225/2020\|EPI_ISL_569388\|20 | ···················· | ···················· | ···················· | ···················· | ···················· | ···················· |
| K | 1001 | hCoV-19/France/PAC-IHU-0226/2020\|EPI_ISL_569389\|20 | ···················· | ···················· | ···················· | ···················· | ···················· | ···················· |
| K | 1002 | hCoV-19/France/PAC-IHU-0227/2020\|EPI_ISL_569390\|20 | ···················· | ···················· | ···················· | ···················· | ···················· | ···················· |
| K | 1003 | hCoV-19/France/PAC-IHU-0228/2020\|EPI_ISL_569391\|20 | ···················· | ···················· | ···················· | ···················· | ···················· | ···················· |
| K | 1004 | hCoV-19/France/PAC-IHU-0229/2020\|EPI_ISL_569392\|20 | ···················· | ···················· | ···················· | ···················· | ···················· | ···················· |
| K | 1005 | hCoV-19/France/PAC-IHU-0230/2020\|EPI_ISL_569393\|20 | ···················· | ···················· | ···················· | ···················· | ···················· | ···················· |
| K | 1006 | hCoV-19/France/PAC-IHU-0231/2020\|EPI_ISL_569394\|20 | ···················· | ···················· | ···················· | ···················· | ···················· | ···················· |
| K | 1007 | hCoV-19/France/PAC-IHU-0232/2020\|EPI_ISL_569395\|20 | ···················· | ···················· | ···················· | ···················· | ···················· | ···················· |
| K | 1008 | hCoV-19/France/PAC-IHU-0234/2020\|EPI_ISL_569397\|20 | ···················· | ···················· | ···················· | ···················· | ···················· | ···················· |
| K | 1009 | hCoV-19/France/PAC-IHU-0235/2020\|EPI_ISL_569398\|20 | ···················· | ···················· | ···················· | ···················· | ···················· | ···················· |
| K | 1010 | hCoV-19/France/PAC-IHU-0236/2020\|EPI_ISL_569399\|20 | ···················· | ···················· | ···················· | ···················· | ···················· | ···················· |
| K | 1011 | hCoV-19/USA/WA-UW-11072/2020\|EPI_ISL_570370\|2020-0 | ···················· | ···················· | ···················· | ···················· | ···················· | ···················· |
| K | 1012 | hCoV-19/USA/WA-UW-11074/2020\|EPI_ISL_570371\|2020-0 | ·T·················· | ···················· | ···················· | ···················· | ···················· | ···················· |
| K | 1013 | hCoV-19/USA/WA-UW-11076/2020\|EPI_ISL_570372\|2020-0 | ···················· | ···················· | ···················· | ···················· | ···················· | ···················· |
| K | 1014 | hCoV-19/USA/WA-UW-11078/2020\|EPI_ISL_570373\|2020-0 | ···················· | ···················· | ···················· | ···················· | ···················· | ···················· |
| K | 1015 | hCoV-19/USA/WA-UW-11098/2020\|EPI_ISL_570375\|2020-0 | ···················· | ···················· | ···················· | ···················· | ···················· | ···················· |
| K | 1016 | hCoV-19/USA/WA-UW-11115/2020\|EPI_ISL_570376\|2020-0 | ···················· | ···················· | ···················· | ···················· | ···················· | ···················· |
| K | 1017 | hCoV-19/USA/OR-UW-11144/2020\|EPI_ISL_570377\|2020-0 | ···················· | ···················· | ···················· | ···················· | ···················· | ···················· |
| K | 1018 | hCoV-19/USA/WA-UW-11165/2020\|EPI_ISL_570378\|2020-0 | ···················· | ···················· | ···················· | ···················· | ···················· | ···················· |
| K | 1019 | hCoV-19/USA/WA-UW-11187/2020\|EPI_ISL_570379\|2020-0 | ···················· | ···················· | ···················· | ···················· | ···················· | ···················· |
| K | 1020 | hCoV-19/USA/WA-UW-11193/2020\|EPI_ISL_570380\|2020-0 | ···················· | ···················· | ···················· | ···················· | ···················· | ···················· |

| | | | | | | | | |
|---|---|---|---|---|---|---|---|---|
| K | 1021 | hCoV-19/USA/WA-UW-11196/2020\|EPI_ISL_570381\|2020-0 | ···················· | ···················· | ····················· | ···················· | ···················· | ···················· |
| K | 1022 | hCoV-19/USA/WA-UW-11259/2020\|EPI_ISL_570382\|2020-0 | ···················· | ···················· | ····················· | ···················· | ···················· | ···················· |
| K | 1023 | hCoV-19/USA/WA-UW-11270/2020\|EPI_ISL_570383\|2020-0 | ···················· | ···················· | ····················· | ···················· | ······T············· | ···················· |
| K | 1024 | hCoV-19/USA/WA-UW-11277/2020\|EPI_ISL_570384\|2020-0 | ···················· | ···················· | ····················· | ···················· | ···················· | ···················· |
| K | 1025 | hCoV-19/USA/UT-UPHL-01413/2020\|EPI_ISL_560841\|2020 | ···················· | ···················· | ····················· | ···················· | ···················· | ···················· |
| K | 1026 | hCoV-19/USA/UT-UPHL-03803/2020\|EPI_ISL_560842\|2020 | ···················· | ···················· | ····················· | ···················· | ···················· | ···················· |
| K | 1027 | hCoV-19/USA/UT-UPHL-03805/2020\|EPI_ISL_560843\|2020 | ···················· | ···················· | ····················· | ···················· | ···················· | ···················· |
| K | 1028 | hCoV-19/USA/UT-UPHL-03938/2020\|EPI_ISL_560844\|2020 | ···················· | ···················· | ····················· | ···················· | ···················· | ···················· |
| K | 1029 | hCoV-19/USA/UT-UPHL-03830/2020\|EPI_ISL_560845\|2020 | ···················· | ···················· | ····················· | ···················· | ···················· | ···················· |
| K | 1030 | hCoV-19/USA/UT-UPHL-01649/2020\|EPI_ISL_560851\|2020 | ···················· | ···················· | ····················· | ···················· | ···················· | ···················· |
| K | 1031 | hCoV-19/USA/UT-UPHL-01650/2020\|EPI_ISL_560852\|2020 | ···················· | ···················· | ····················· | ···················· | ···················· | ···················· |
| K | 1032 | hCoV-19/USA/UT-UPHL-01651/2020\|EPI_ISL_560853\|2020 | ···················· | ···················· | ····················· | ···················· | ···················· | ···················· |
| K | 1033 | hCoV-19/USA/UT-UPHL-01656/2020\|EPI_ISL_560856\|2020 | ···················· | ···················· | ····················· | ···················· | ···················· | ···················· |
| K | 1034 | hCoV-19/USA/UT-UPHL-01657/2020\|EPI_ISL_560857\|2020 | ···················· | ···················· | ····················· | ···················· | ···················· | ···················· |
| K | 1035 | hCoV-19/USA/UT-UPHL-01658/2020\|EPI_ISL_560858\|2020 | ···················· | ···················· | ····················· | ···················· | ···················· | ···················· |
| K | 1036 | hCoV-19/USA/UT-UPHL-01659/2020\|EPI_ISL_560859\|2020 | ···················· | ···················· | ····················· | ···················· | ···················· | ···················· |
| K | 1037 | hCoV-19/USA/UT-UPHL-01660/2020\|EPI_ISL_560860\|2020 | ···················· | ···················· | ····················· | ···················· | ···················· | ···················· |
| K | 1038 | hCoV-19/USA/UT-UPHL-03843/2020\|EPI_ISL_560861\|2020 | ···················· | ···················· | ····················· | ···················· | ···················· | ···················· |
| K | 1039 | hCoV-19/USA/UT-UPHL-05172/2020\|EPI_ISL_560862\|2020 | ···················· | ···················· | ····················· | ···················· | ···················· | ···················· |
| K | 1040 | hCoV-19/USA/UT-UPHL-05175/2020\|EPI_ISL_560863\|2020 | ···················· | ···················· | ····················· | ···················· | ···················· | ···················· |
| K | 1041 | hCoV-19/USA/UT-UPHL-05209/2020\|EPI_ISL_560865\|2020 | ···················· | ···················· | ····················· | ···················· | ···················· | ···················· |
| K | 1042 | hCoV-19/USA/UT-UPHL-05304/2020\|EPI_ISL_560870\|2020 | ···················· | ···················· | ····················· | ···················· | ···················· | ···················· |
| K | 1043 | hCoV-19/USA/UT-UPHL-05411/2020\|EPI_ISL_560875\|2020 | ···················· | ···················· | ····················· | ···················· | ···················· | ···················· |
| K | 1044 | hCoV-19/USA/UT-UPHL-05426/2020\|EPI_ISL_560878\|2020 | ···················· | ···················· | ····················· | ···················· | ···················· | ···················· |
| K | 1045 | hCoV-19/USA/UT-UPHL-05443/2020\|EPI_ISL_560880\|2020 | ···················· | ···················· | ····················· | ···················· | ···················· | ···················· |
| K | 1046 | hCoV-19/USA/UT-UPHL-05479/2020\|EPI_ISL_560882\|2020 | ···················· | ···················· | ····················· | ···················· | ···················· | ···················· |
| K | 1047 | hCoV-19/USA/UT-UPHL-05516/2020\|EPI_ISL_560884\|2020 | ···················· | ···················· | ····················· | ···················· | ···················· | ···················· |
| K | 1048 | hCoV-19/USA/UT-UPHL-05531/2020\|EPI_ISL_560886\|2020 | ···················· | ···················· | ····················· | ···················· | ···················· | ···················· |
| K | 1049 | hCoV-19/USA/UT-UPHL-05537/2020\|EPI_ISL_560888\|2020 | ···················· | ···················· | ····················· | ···················· | ···················· | ···················· |
| K | 1050 | hCoV-19/USA/UT-UPHL-05549/2020\|EPI_ISL_560889\|2020 | ···················· | ···················· | ····················· | ···················· | ···················· | ···················· |
| K | 1051 | hCoV-19/USA/UT-UPHL-05578/2020\|EPI_ISL_560892\|2020 | ···················· | ···················· | ····················· | ···················· | ···················· | ···················· |
| K | 1052 | hCoV-19/USA/UT-UPHL-05593/2020\|EPI_ISL_560894\|2020 | ···················· | ···················· | ····················· | ···················· | ···················· | ···················· |
| K | 1053 | hCoV-19/USA/UT-UPHL-03046/2020\|EPI_ISL_560897\|2020 | ···················· | ·A·················· | ····················· | ···················· | ···················· | ···················· |
| K | 1054 | hCoV-19/USA/UT-UPHL-03064/2020\|EPI_ISL_560899\|2020 | ···················· | ···················· | ····················· | ···················· | ···················· | ···················· |
| K | 1055 | hCoV-19/Australia/VIC10711/2020\|EPI_ISL_563048\|202 | ···················· | ···················· | ····················· | ···················· | ···················· | ···················· |
| K | 1056 | hCoV-19/Australia/VIC10712/2020\|EPI_ISL_563049\|202 | ···················· | ···················· | ····················· | ···················· | ···················· | ···················· |
| K | 1057 | hCoV-19/Australia/VIC10718/2020\|EPI_ISL_563051\|202 | ···················· | ···················· | ····················· | ···················· | ···················· | ···················· |
| K | 1058 | hCoV-19/Australia/VIC10719/2020\|EPI_ISL_563052\|202 | ···················· | ···················· | ····················· | ···················· | ···················· | ···················· |
| K | 1059 | hCoV-19/Australia/VIC10720/2020\|EPI_ISL_563053\|202 | ···················· | ···················· | ····················· | ···················· | ···················· | ···················· |
| K | 1060 | hCoV-19/Australia/VIC10721/2020\|EPI_ISL_563054\|202 | ···················· | ···················· | ····················· | ···················· | ···················· | ···················· |
| K | 1061 | hCoV-19/Australia/VIC10723/2020\|EPI_ISL_563055\|202 | ···················· | ···················· | ····················· | ···················· | ···················· | ···················· |
| K | 1062 | hCoV-19/Australia/VIC10725/2020\|EPI_ISL_563056\|202 | ···················· | ···················· | ····················· | ···················· | ···················· | ···················· |
| K | 1063 | hCoV-19/Australia/VIC10726/2020\|EPI_ISL_563057\|202 | ···················· | ···················· | ····················· | ···················· | ···················· | ···················· |
| K | 1064 | hCoV-19/Australia/VIC10727/2020\|EPI_ISL_563058\|202 | ···················· | ···················· | ····················· | ···················· | ···················· | ···················· |
| K | 1065 | hCoV-19/Australia/VIC7393/2020\|EPI_ISL_564379\|2020 | ···················· | ···················· | ····················· | ···················· | ···················· | ···················· |
| K | 1066 | hCoV-19/Australia/VIC7394/2020\|EPI_ISL_564380\|2020 | ···················· | ···················· | ····················· | ···················· | ···················· | ···················· |
| K | 1067 | hCoV-19/Australia/VIC7396/2020\|EPI_ISL_564381\|2020 | ···················· | ···················· | ····················· | ···················· | ···················· | ···················· |
| K | 1068 | hCoV-19/Australia/VIC7398/2020\|EPI_ISL_564382\|2020 | ···················· | ···················· | ····················· | ···················· | ···················· | ···················· |
| K | 1069 | hCoV-19/Australia/VIC7399/2020\|EPI_ISL_564383\|2020 | ···················· | ···················· | ····················· | ···················· | ···················· | ···················· |
| K | 1070 | hCoV-19/Australia/VIC7400/2020\|EPI_ISL_564384\|2020 | ···················· | ···················· | ····················· | ···················· | ···················· | ···················· |
| K | 1071 | hCoV-19/Australia/VIC7403/2020\|EPI_ISL_564385\|2020 | ···················· | ···················· | ····················· | ···················· | ···················· | ···················· |
| K | 1072 | hCoV-19/Australia/VIC7404/2020\|EPI_ISL_564386\|2020 | ···················· | ·R·················· | ····················· | ···················· | ···················· | ···················· |
| K | 1073 | hCoV-19/Australia/VIC7405/2020\|EPI_ISL_564387\|2020 | ···················· | ···················· | ····················· | ···················· | ···················· | ···················· |
| K | 1074 | hCoV-19/Australia/VIC7406/2020\|EPI_ISL_564388\|2020 | ···················· | ···················· | ····················· | ···················· | ···················· | ···················· |
| K | 1075 | hCoV-19/Australia/VIC7409/2020\|EPI_ISL_564389\|2020 | ···················· | ···················· | ····················· | ···················· | ···················· | ···················· |
| K | 1076 | hCoV-19/Australia/VIC7562/2020\|EPI_ISL_564500\|2020 | ···················· | ···················· | ····················· | ···················· | ···················· | ···················· |
| K | 1077 | hCoV-19/Australia/VIC7563/2020\|EPI_ISL_564501\|2020 | ···················· | ···················· | ····················· | ···················· | ···················· | ···················· |
| K | 1078 | hCoV-19/Australia/VIC7565/2020\|EPI_ISL_564502\|2020 | ···················· | ···················· | ····················· | ···················· | ···················· | ···················· |
| K | 1079 | hCoV-19/Australia/VIC7567/2020\|EPI_ISL_564503\|2020 | ···················· | ···················· | ····················· | ···················· | ···················· | ···················· |
| K | 1080 | hCoV-19/Australia/VIC7569/2020\|EPI_ISL_564504\|2020 | ···················· | ···················· | ····················· | ···················· | ···················· | ···················· |
| K | 1081 | hCoV-19/Australia/VIC7570/2020\|EPI_ISL_564505\|2020 | ···················· | ···················· | ····················· | ···················· | ···················· | ···················· |
| K | 1082 | hCoV-19/Australia/VIC7571/2020\|EPI_ISL_564506\|2020 | ···················· | ···················· | ····················· | ···················· | ···················· | ···················· |
| K | 1083 | hCoV-19/Australia/VIC7573/2020\|EPI_ISL_564507\|2020 | ···················· | ···················· | ····················· | ···················· | ···················· | ···················· |
| K | 1084 | hCoV-19/Australia/VIC7576/2020\|EPI_ISL_564508\|2020 | ···················· | ···················· | ····················· | ···················· | ···················· | ···················· |
| K | 1085 | hCoV-19/Australia/VIC7578/2020\|EPI_ISL_564509\|2020 | ···················· | ···················· | ····················· | ···················· | ···················· | ···················· |
| K | 1086 | hCoV-19/Australia/VIC7580/2020\|EPI_ISL_564510\|2020 | ···················· | ···················· | ····················· | ···················· | ···················· | ···················· |
| K | 1087 | hCoV-19/Australia/VIC7581/2020\|EPI_ISL_564511\|2020 | ···················· | ···················· | ····················· | ···················· | ···················· | ···················· |
| K | 1088 | hCoV-19/Australia/VIC7582/2020\|EPI_ISL_564512\|2020 | ···················· | ···················· | ····················· | ···················· | ···················· | ···················· |
| K | 1089 | hCoV-19/Australia/VIC7583/2020\|EPI_ISL_564513\|2020 | ···················· | ···················· | ····················· | ···················· | ···················· | ···················· |
| K | 1090 | hCoV-19/Australia/VIC7586/2020\|EPI_ISL_564514\|2020 | ···················· | ···················· | ····················· | ···················· | ···················· | ···················· |
| K | 1091 | hCoV-19/Australia/VIC7589/2020\|EPI_ISL_564515\|2020 | ···················· | ···················· | ····················· | ···················· | ···················· | ···················· |
| K | 1092 | hCoV-19/Australia/VIC7590/2020\|EPI_ISL_564516\|2020 | ···················· | ···················· | ····················· | ···················· | ···················· | ···················· |
| K | 1093 | hCoV-19/Australia/VIC7591/2020\|EPI_ISL_564517\|2020 | ···················· | ···················· | ····················· | ···················· | ···················· | ···················· |
| K | 1094 | hCoV-19/Australia/VIC7592/2020\|EPI_ISL_564518\|2020 | ···················· | ···················· | ····················· | ···················· | ···················· | ···················· |
| K | 1095 | hCoV-19/Australia/VIC7594/2020\|EPI_ISL_564519\|2020 | ···················· | ···················· | ····················· | ···················· | ···················· | ···················· |
| K | 1096 | hCoV-19/Australia/VIC7596/2020\|EPI_ISL_564520\|2020 | ···················· | ···················· | ····················· | ···················· | ···················· | ···················· |
| K | 1097 | hCoV-19/Australia/VIC7597/2020\|EPI_ISL_564521\|2020 | ···················· | ···················· | ····················· | ···················· | ···················· | ···················· |
| K | 1098 | hCoV-19/Australia/VIC9952/2020\|EPI_ISL_565800\|2020 | ···················· | ···················· | ····················· | ···················· | ···················· | ···················· |
| K | 1099 | hCoV-19/Australia/VIC9955/2020\|EPI_ISL_565801\|2020 | ···················· | ···················· | ····················· | ···················· | ···················· | ···················· |
| K | 1100 | hCoV-19/Australia/VIC9956/2020\|EPI_ISL_565802\|2020 | ···················· | ···················· | ····················· | ···················· | ···················· | ···················· |

| | | | | | | | |
|---|---|---|---|---|---|---|---|
| K | 1101 hCoV-19/Australia/VIC9957/2020\|EPI_ISL_565803\|2020 | .................... | .................... | .................... | .................... | .................... | .................... |
| K | 1102 hCoV-19/Australia/VIC9958/2020\|EPI_ISL_565804\|2020 | .................... | .................... | .................... | .................... | .................... | .................... |
| K | 1103 hCoV-19/Australia/VIC9959/2020\|EPI_ISL_565805\|2020 | .................... | .................... | .................... | .................... | .................... | .................... |
| K | 1104 hCoV-19/Australia/VIC9961/2020\|EPI_ISL_565806\|2020 | .................... | .................... | .................... | .................... | .................... | .................... |
| K | 1105 hCoV-19/Australia/VIC9962/2020\|EPI_ISL_565807\|2020 | .................... | .................... | .................... | .................... | .................... | .................... |
| K | 1106 hCoV-19/Australia/VIC9964/2020\|EPI_ISL_565808\|2020 | .................... | .................... | .................... | .................... | .................... | .................... |
| K | 1107 hCoV-19/Australia/VIC9965/2020\|EPI_ISL_565809\|2020 | .................... | .................... | .................... | .................... | .................... | .................... |
| K | 1108 hCoV-19/Australia/VIC9966/2020\|EPI_ISL_565810\|2020 | .................... | .................... | .................... | .................... | .................... | .................... |
| K | 1109 hCoV-19/Australia/VIC9967/2020\|EPI_ISL_565811\|2020 | .................... | .................... | .................... | .................... | .................... | .................... |
| K | 1110 hCoV-19/Australia/VIC9968/2020\|EPI_ISL_565812\|2020 | .................... | .................... | .................... | .................... | .................... | .................... |
| K | 1111 hCoV-19/Australia/VIC9969/2020\|EPI_ISL_565813\|2020 | .................... | .................... | .................... | .................... | .................... | .................... |
| K | 1112 hCoV-19/Australia/VIC9971/2020\|EPI_ISL_565814\|2020 | .................... | .................... | .................... | .................... | .................... | .................... |
| K | 1113 hCoV-19/Australia/VIC9972/2020\|EPI_ISL_565815\|2020 | .................... | .................... | .................... | .................... | .................... | .................... |
| K | 1114 hCoV-19/Australia/VIC9973/2020\|EPI_ISL_565816\|2020 | .................... | .................... | .................... | .................... | .................... | .................... |
| K | 1115 hCoV-19/Australia/VIC9974/2020\|EPI_ISL_565817\|2020 | .................... | .................... | .................... | .................... | .................... | .................... |
| K | 1116 hCoV-19/Australia/VIC9975/2020\|EPI_ISL_565818\|2020 | .................... | .................... | .................... | .................... | .................... | .................... |
| K | 1117 hCoV-19/Australia/VIC9976/2020\|EPI_ISL_565819\|2020 | .................... | .................... | .................... | .................... | .................... | .................... |
| K | 1118 hCoV-19/Australia/VIC9977/2020\|EPI_ISL_565820\|2020 | .................... | .................... | .................... | .................... | .................... | .................... |
| K | 1119 hCoV-19/Australia/VIC9978/2020\|EPI_ISL_565821\|2020 | .................... | .................... | .................... | .................... | .................... | .................... |
| K | 1120 hCoV-19/Australia/VIC9979/2020\|EPI_ISL_565822\|2020 | .................... | .................... | .................... | .................... | .................... | .................... |
| K | 1121 hCoV-19/Australia/VIC9985/2020\|EPI_ISL_565823\|2020 | .................... | .................... | .................... | .................... | .................... | .................... |
| K | 1122 hCoV-19/Australia/VIC9986/2020\|EPI_ISL_565824\|2020 | .................... | .................... | .................... | .................... | .................... | .................... |
| K | 1123 hCoV-19/Australia/VIC9992/2020\|EPI_ISL_565825\|2020 | .................... | .................... | .................... | .................... | .................... | .................... |
| K | 1124 hCoV-19/Australia/VIC9993/2020\|EPI_ISL_565826\|2020 | .................... | .................... | .................... | .................... | .................... | .................... |
| K | 1125 hCoV-19/Australia/VIC9995/2020\|EPI_ISL_565827\|2020 | .................... | .................... | .................... | .................... | .................... | .................... |
| K | 1126 hCoV-19/Australia/VIC9997/2020\|EPI_ISL_565828\|2020 | .................... | .................... | .................... | .................... | .................... | .................... |
| K | 1127 hCoV-19/Australia/VIC9999/2020\|EPI_ISL_565829\|2020 | .................... | .................... | .................... | .................... | .................... | .................... |
| K | 1128 hCoV-19/USA/DE-DHSS-F917719/2020\|EPI_ISL_565830\|20 | .................... | .................... | .................... | .................... | .................... | .................... |
| K | 1129 hCoV-19/USA/DE-DHSS-F917751/2020\|EPI_ISL_565832\|20 | .................... | .................... | .................... | .................... | .................... | .................... |
| K | 1130 hCoV-19/USA/MI-MDHHS-SC21794/2020\|EPI_ISL_565837\|2 | .................... | .................... | .................... | .................... | .................... | .................... |
| K | 1131 hCoV-19/USA/MI-MDHHS-SC21795/2020\|EPI_ISL_565838\|2 | .................... | .................... | .................... | .................... | .................... | .................... |
| K | 1132 hCoV-19/USA/MI-MDHHS-SC21800/2020\|EPI_ISL_565843\|2 | .................... | .................... | .................... | .................... | .................... | .................... |
| K | 1133 hCoV-19/USA/MI-MDHHS-SC21803/2020\|EPI_ISL_565846\|2 | .................... | .................... | .................... | .................... | .................... | .................... |
| K | 1134 hCoV-19/USA/MI-MDHHS-SC21806/2020\|EPI_ISL_565849\|2 | .................... | .................... | .................... | .................... | .................... | .................... |
| K | 1135 hCoV-19/England/ALDP-9BE81F/2020\|EPI_ISL_568050\|20 | .................... | .................... | .................... | .................... | .................... | .................... |
| K | 1136 hCoV-19/England/ALDP-9BC714/2020\|EPI_ISL_568051\|20 | .................... | .................... | .................... | .................... | .................... | .................... |
| K | 1137 hCoV-19/England/ALDP-9BED2F/2020\|EPI_ISL_568052\|20 | .................... | .................... | .................... | .................... | .................... | .................... |
| K | 1138 hCoV-19/England/ALDP-9BEA82/2020\|EPI_ISL_568053\|20 | .................... | .................... | .................... | .................... | .................... | .................... |
| K | 1139 hCoV-19/England/ALDP-9BDB26/2020\|EPI_ISL_568054\|20 | .................... | .................... | .................... | .................... | .................... | .................... |
| K | 1140 hCoV-19/England/ALDP-9BE299/2020\|EPI_ISL_568055\|20 | .................... | .................... | .................... | .................... | .................... | .................... |
| K | 1141 hCoV-19/England/ALDP-9BEC13/2020\|EPI_ISL_568070\|20 | .................... | .................... | .................... | .................... | .................... | .................... |
| K | 1142 hCoV-19/England/ALDP-9BE132/2020\|EPI_ISL_568071\|20 | .................... | .................... | .................... | .................... | .................... | .................... |
| K | 1143 hCoV-19/England/ALDP-9BDFC0/2020\|EPI_ISL_568072\|20 | .................... | .................... | .................... | .................... | .................... | .................... |
| K | 1144 hCoV-19/England/ALDP-9BEDB6/2020\|EPI_ISL_568073\|20 | .................... | .................... | .................... | .................... | .................... | .................... |
| K | 1145 hCoV-19/England/ALDP-9BE387/2020\|EPI_ISL_568074\|20 | .................... | .................... | .................... | .................... | .................... | .................... |
| K | 1146 hCoV-19/England/ALDP-9BDADE/2020\|EPI_ISL_568075\|20 | .................... | .................... | .................... | .................... | .................... | .................... |
| K | 1147 hCoV-19/England/ALDP-9BE0DB/2020\|EPI_ISL_568076\|20 | .................... | .................... | .................... | .................... | .................... | .................... |
| K | 1148 hCoV-19/England/ALDP-9BE83D/2020\|EPI_ISL_568079\|20 | .................... | .................... | .................... | .................... | .................... | .................... |
| K | 1149 hCoV-19/England/ALDP-9BE4FD/2020\|EPI_ISL_568080\|20 | .................... | .................... | .................... | .................... | .................... | .................... |
| K | 1150 hCoV-19/England/ALDP-9BE4B1/2020\|EPI_ISL_568081\|20 | .................... | .................... | .................... | .................... | .................... | .................... |
| K | 1151 hCoV-19/England/ALDP-9BDD3F/2020\|EPI_ISL_568082\|20 | .................... | .................... | .................... | .................... | .................... | .................... |
| K | 1152 hCoV-19/England/ALDP-9BEDC5/2020\|EPI_ISL_568083\|20 | .................... | .................... | .................... | .................... | .................... | .................... |
| K | 1153 hCoV-19/England/ALDP-9BEB61/2020\|EPI_ISL_568084\|20 | .................... | .................... | .................... | .................... | .................... | .................... |
| K | 1154 hCoV-19/England/ALDP-9BEC04/2020\|EPI_ISL_568085\|20 | .................... | .................... | .................... | .................... | .................... | .................... |
| K | 1155 hCoV-19/England/ALDP-9BEC31/2020\|EPI_ISL_568086\|20 | .................... | .................... | .................... | .................... | .................... | .................... |
| K | 1156 hCoV-19/England/ALDP-9BE69D/2020\|EPI_ISL_568087\|20 | .................... | .................... | .................... | .................... | .................... | .................... |
| K | 1157 hCoV-19/England/ALDP-9BE31E/2020\|EPI_ISL_568088\|20 | .................... | .................... | .................... | .................... | .................... | .................... |
| K | 1158 hCoV-19/England/ALDP-9BDF66/2020\|EPI_ISL_568089\|20 | .................... | .................... | .................... | .................... | .................... | .................... |
| K | 1159 hCoV-19/England/ALDP-9BDCAB/2020\|EPI_ISL_568090\|20 | .................... | .................... | .................... | .................... | .................... | .................... |
| K | 1160 hCoV-19/England/ALDP-9BE493/2020\|EPI_ISL_568091\|20 | .................... | .................... | .................... | .................... | .................... | .................... |
| K | 1161 hCoV-19/England/ALDP-9BE7D6/2020\|EPI_ISL_568092\|20 | .................... | .................... | .................... | .................... | .................... | .................... |
| K | 1162 hCoV-19/England/ALDP-9BE9D0/2020\|EPI_ISL_568093\|20 | .................... | .................... | .................... | .................... | .................... | .................... |
| K | 1163 hCoV-19/England/ALDP-9BE369/2020\|EPI_ISL_568094\|20 | .................... | .................... | .................... | .................... | .................... | .................... |
| K | 1164 hCoV-19/England/ALDP-9BDEC3/2020\|EPI_ISL_568095\|20 | .................... | .................... | .................... | .................... | .................... | .................... |
| K | 1165 hCoV-19/England/ALDP-9BE396/2020\|EPI_ISL_568097\|20 | .................... | .................... | .................... | .................... | .................... | .................... |
| K | 1166 hCoV-19/England/ALDP-9BDEB4/2020\|EPI_ISL_568098\|20 | .................... | .A.................. | .................... | .................... | .................... | ..A................. |
| K | 1167 hCoV-19/England/ALDP-9BE958/2020\|EPI_ISL_568099\|20 | .................... | .A.................. | .................... | .................... | .................... | ..A................. |
| K | 1168 hCoV-19/France/PAC-IHU-1567/2020\|EPI_ISL_569200\|20 | .................... | .................... | .................... | .................... | .................... | .................... |
| K | 1169 hCoV-19/France/PAC-IHU-1568/2020\|EPI_ISL_569201\|20 | .................... | .................... | .................... | .................... | .................... | .................... |
| K | 1170 hCoV-19/France/PAC-IHU-1569/2020\|EPI_ISL_569202\|20 | .................... | .................... | .................... | .................... | .................... | .................... |
| K | 1171 hCoV-19/France/PAC-IHU-1570/2020\|EPI_ISL_569203\|20 | .................... | .................... | .................... | .................... | .................... | .................... |
| K | 1172 hCoV-19/France/PAC-IHU-1571/2020\|EPI_ISL_569204\|20 | .................... | .................... | .................... | .................... | .................... | .................... |
| K | 1173 hCoV-19/France/PAC-IHU-1572/2020\|EPI_ISL_569205\|20 | .................... | .................... | .................... | .................... | .................... | .................... |
| K | 1174 hCoV-19/France/PAC-IHU-1574/2020\|EPI_ISL_569207\|20 | .................... | .................... | .................... | .................... | .................... | .................... |
| K | 1175 hCoV-19/France/PAC-IHU-1575/2020\|EPI_ISL_569208\|20 | .................... | .................... | .................... | .................... | .................... | .................... |
| K | 1176 hCoV-19/France/PAC-IHU-1581/2020\|EPI_ISL_569210\|20 | .................... | .................... | .................... | .................... | .................... | .................... |
| K | 1177 hCoV-19/France/PAC-IHU-1582/2020\|EPI_ISL_569211\|20 | .................... | .................... | .................... | .................... | .................... | .................... |
| K | 1178 hCoV-19/France/PAC-IHU-1585/2020\|EPI_ISL_569214\|20 | .................... | .................... | .................... | .................... | .................... | .................... |
| K | 1179 hCoV-19/France/PAC-IHU-1587/2020\|EPI_ISL_569216\|20 | .................... | .................... | .................... | .................... | .................... | .................... |
| K | 1180 hCoV-19/France/PAC-IHU-1588/2020\|EPI_ISL_569217\|20 | .................... | .................... | .................... | .................... | .................... | .................... |

| | | | | | | | | |
|---|---|---|---|---|---|---|---|---|
| K | 1181 | hCoV-19/France/PAC-IHU-1590/2020\|EPI_ISL_569219\|20 | .................... | .................... | .................... | .................... | .................... | .................... |
| K | 1182 | hCoV-19/France/PAC-IHU-1591/2020\|EPI_ISL_569220\|20 | .................... | .................... | .................... | .................... | .................... | .................... |
| K | 1183 | hCoV-19/France/PAC-IHU-1592/2020\|EPI_ISL_569221\|20 | .................... | .................... | .................... | .................... | .................... | .................... |
| K | 1184 | hCoV-19/France/PAC-IHU-1597/2020\|EPI_ISL_569222\|20 | .................... | .................... | .................... | .................... | .................... | .................... |
| K | 1185 | hCoV-19/France/PAC-IHU-1599/2020\|EPI_ISL_569223\|20 | .................... | .................... | .................... | .................... | .................... | .................... |
| K | 1186 | hCoV-19/France/PAC-IHU-1607/2020\|EPI_ISL_569225\|20 | .................... | .................... | .................... | .................... | .................... | .................... |
| K | 1187 | hCoV-19/France/PAC-IHU-1609/2020\|EPI_ISL_569226\|20 | .................... | .................... | .................... | .................... | .................... | .................... |
| K | 1188 | hCoV-19/France/PAC-IHU-1610/2020\|EPI_ISL_569227\|20 | .................... | .................... | .................... | .................... | .................... | .................... |
| K | 1189 | hCoV-19/France/PAC-IHU-1615/2020\|EPI_ISL_569228\|20 | .................... | .................... | .................... | .................... | .................... | .................... |
| K | 1190 | hCoV-19/France/PAC-IHU-1618/2020\|EPI_ISL_569229\|20 | .................... | .................... | .................... | .................... | .................... | .................... |
| K | 1191 | hCoV-19/France/PAC-IHU-1620/2020\|EPI_ISL_569230\|20 | .................... | .................... | .................... | .................... | .................... | .................... |
| K | 1192 | hCoV-19/France/PAC-IHU-1622/2020\|EPI_ISL_569231\|20 | .................... | .................... | .................... | .................... | .................... | .................... |
| K | 1193 | hCoV-19/France/PAC-IHU-1623/2020\|EPI_ISL_569232\|20 | .................... | .................... | .................... | .................... | .................... | .................... |
| K | 1194 | hCoV-19/France/PAC-IHU-1624/2020\|EPI_ISL_569233\|20 | .................... | .................... | .................... | .................... | .................... | .................... |
| K | 1195 | hCoV-19/France/PAC-IHU-1625/2020\|EPI_ISL_569234\|20 | .................... | .................... | .................... | .................... | .................... | .................... |
| K | 1196 | hCoV-19/France/PAC-IHU-1626/2020\|EPI_ISL_569235\|20 | .................... | .................... | .................... | .................... | .................... | .................... |
| K | 1197 | hCoV-19/France/PAC-IHU-1627/2020\|EPI_ISL_569236\|20 | .................... | .................... | .................... | .................... | .................... | .................... |
| K | 1198 | hCoV-19/France/PAC-IHU-1628/2020\|EPI_ISL_569237\|20 | .................... | .................... | .................... | .................... | .................... | .................... |
| K | 1199 | hCoV-19/France/PAC-IHU-1629/2020\|EPI_ISL_569238\|20 | .................... | .................... | .................... | .................... | .................... | .................... |
| K | 1200 | hCoV-19/France/PAC-IHU-1630/2020\|EPI_ISL_569239\|20 | .................... | .................... | .................... | .................... | .................... | .................... |
| K | 1201 | hCoV-19/France/PAC-IHU-1631/2020\|EPI_ISL_569240\|20 | .................... | .................... | .................... | .................... | .................... | .................... |
| K | 1202 | hCoV-19/France/PAC-IHU-1632/2020\|EPI_ISL_569241\|20 | .................... | .................... | .................... | .................... | .................... | .................... |
| K | 1203 | hCoV-19/France/PAC-IHU-0012/2020\|EPI_ISL_569243\|20 | .................... | .................... | .................... | .................... | .................... | .................... |
| K | 1204 | hCoV-19/France/PAC-IHU-0014/2020\|EPI_ISL_569244\|20 | .................... | .................... | .................... | .................... | .................... | .................... |
| K | 1205 | hCoV-19/France/PAC-IHU-0017/2020\|EPI_ISL_569246\|20 | .................... | .................... | .................... | .................... | .................... | .................... |
| K | 1206 | hCoV-19/France/PAC-IHU-0018/2020\|EPI_ISL_569247\|20 | .................... | .................... | .................... | .................... | .................... | .................... |
| K | 1207 | hCoV-19/France/PAC-IHU-0019/2020\|EPI_ISL_569248\|20 | .................... | .................... | .................... | .................... | .................... | .................... |
| K | 1208 | hCoV-19/France/PAC-IHU-0020/2020\|EPI_ISL_569249\|20 | .................... | .................... | .................... | .................... | .................... | .................... |
| K | 1209 | hCoV-19/France/PAC-IHU-0021/2020\|EPI_ISL_569250\|20 | .................... | .................... | .................... | .................... | .................... | .................... |
| K | 1210 | hCoV-19/France/PAC-IHU-0022/2020\|EPI_ISL_569251\|20 | .................... | .................... | .................... | .................... | .................... | .................... |
| K | 1211 | hCoV-19/France/PAC-IHU-0024/2020\|EPI_ISL_569252\|20 | .................... | .................... | .................... | .................... | .................... | .................... |
| K | 1212 | hCoV-19/France/PAC-IHU-0026/2020\|EPI_ISL_569253\|20 | .................... | .................... | .................... | .................... | .................... | .................... |
| K | 1213 | hCoV-19/France/PAC-IHU-0028/2020\|EPI_ISL_569254\|20 | .................... | .................... | .................... | .................... | .................... | .................... |
| K | 1214 | hCoV-19/France/PAC-IHU-0029/2020\|EPI_ISL_569255\|20 | .................... | .................... | .................... | .................... | .................... | .................... |
| K | 1215 | hCoV-19/France/PAC-IHU-0030/2020\|EPI_ISL_569256\|20 | .................... | .................... | .................... | .................... | .................... | .................... |
| K | 1216 | hCoV-19/France/PAC-IHU-0031/2020\|EPI_ISL_569257\|20 | .................... | .................... | .................... | .................... | .................... | .................... |
| K | 1217 | hCoV-19/France/PAC-IHU-0032/2020\|EPI_ISL_569258\|20 | .................... | .................... | .................... | .................... | .................... | .................... |
| K | 1218 | hCoV-19/France/PAC-IHU-0033/2020\|EPI_ISL_569259\|20 | .................... | .................... | .................... | .................... | .................... | .................... |
| K | 1219 | hCoV-19/France/PAC-IHU-0215/2020\|EPI_ISL_569380\|20 | .................... | .................... | .................... | .................... | .................... | .................... |
| K | 1220 | hCoV-19/France/PAC-IHU-0216/2020\|EPI_ISL_569381\|20 | .................... | .................... | .................... | .................... | .................... | .................... |
| K | 1221 | hCoV-19/France/PAC-IHU-0217/2020\|EPI_ISL_569382\|20 | .................... | .................... | .................... | .................... | .................... | .................... |
| K | 1222 | hCoV-19/France/PAC-IHU-0218/2020\|EPI_ISL_569383\|20 | .................... | .................... | .................... | .................... | .................... | .................... |
| K | 1223 | hCoV-19/France/PAC-IHU-0219/2020\|EPI_ISL_569384\|20 | .................... | .................... | .................... | .................... | .................... | .................... |
| K | 1224 | hCoV-19/France/PAC-IHU-0222/2020\|EPI_ISL_569386\|20 | .................... | .................... | .................... | .................... | .................... | .................... |
| K | 1225 | hCoV-19/USA/WA-UW-8165/2020\|EPI_ISL_570200\|2020-04 | .................... | .................... | .................... | .................... | .................... | .................... |
| K | 1226 | hCoV-19/USA/WA-UW-831/2020\|EPI_ISL_570201\|2020-03- | .................... | .................... | .................... | .................... | .................... | .................... |
| K | 1227 | hCoV-19/USA/WA-UW-8317/2020\|EPI_ISL_570202\|2020-04 | .................... | .................... | .................... | .................... | .................... | .................... |
| K | 1228 | hCoV-19/USA/WA-UW-8330/2020\|EPI_ISL_570203\|2020-04 | .................... | .................... | .................... | .................... | .................... | .................... |
| K | 1229 | hCoV-19/USA/WA-UW-8418/2020\|EPI_ISL_570205\|2020-05 | .................... | .................... | .................... | .................... | .................... | .................... |
| K | 1230 | hCoV-19/USA/WA-UW-8421/2020\|EPI_ISL_570206\|2020-05 | .................... | .................... | .................... | .................... | .................... | .................... |
| K | 1231 | hCoV-19/USA/WA-UW-8422/2020\|EPI_ISL_570207\|2020-05 | .................... | .................... | .................... | .................... | .................... | .................... |
| K | 1232 | hCoV-19/USA/WA-UW-8423/2020\|EPI_ISL_570208\|2020-05 | .................... | .................... | .................... | .................... | .................... | .................... |
| K | 1233 | hCoV-19/USA/WA-UW-8511/2020\|EPI_ISL_570211\|2020-05 | .................... | .................... | .................... | .................... | .................... | .................... |
| K | 1234 | hCoV-19/USA/WA-UW-8526/2020\|EPI_ISL_570213\|2020-05 | .................... | .................... | .................... | .................... | .................... | .................... |
| K | 1235 | hCoV-19/USA/WA-UW-8550/2020\|EPI_ISL_570214\|2020-05 | .................... | .................... | .................... | .................... | .................... | .................... |
| K | 1236 | hCoV-19/USA/WA-UW-8636/2020\|EPI_ISL_570215\|2020-05 | .................... | .................... | .................... | .................... | .................... | .................... |
| K | 1237 | hCoV-19/USA/WA-UW-8698/2020\|EPI_ISL_570217\|2020-05 | .................... | .................... | .................... | .................... | .................... | .................... |
| K | 1238 | hCoV-19/USA/WA-UW-8813/2020\|EPI_ISL_570219\|2020-05 | .................... | .................... | .................... | .................... | .................... | .................... |
| K | 1239 | hCoV-19/USA/WA-UW-8831/2020\|EPI_ISL_570221\|2020-05 | .................... | .................... | .................... | .................... | .................... | .................... |
| K | 1240 | hCoV-19/USA/WA-UW-8851/2020\|EPI_ISL_570222\|2020-05 | .................... | .................... | .................... | .................... | .................... | .................... |
| K | 1241 | hCoV-19/USA/WA-UW-8872/2020\|EPI_ISL_570223\|2020-05 | .................... | .................... | .................... | .................... | .................... | .................... |
| K | 1242 | hCoV-19/USA/WA-UW-8873/2020\|EPI_ISL_570224\|2020-05 | .................... | .................... | .................... | .................... | .................... | .................... |
| K | 1243 | hCoV-19/USA/WA-UW-9014/2020\|EPI_ISL_570225\|2020-05 | .................... | .................... | .................... | .................... | .................... | .................... |
| K | 1244 | hCoV-19/USA/WA-UW-9016/2020\|EPI_ISL_570226\|2020-05 | .................... | .................... | .................... | .................... | .................... | .................... |
| K | 1245 | hCoV-19/USA/WA-UW-9068/2020\|EPI_ISL_570227\|2020-05 | .................... | .................... | .................... | .................... | .................... | .................... |
| K | 1246 | hCoV-19/USA/WA-UW-9069/2020\|EPI_ISL_570228\|2020-05 | .................... | .................... | .................... | .................... | .................... | .................... |
| K | 1247 | hCoV-19/USA/WA-UW-9190/2020\|EPI_ISL_570229\|2020-05 | .................... | .................... | .................... | .................... | .................... | .................... |
| K | 1248 | hCoV-19/USA/WA-UW-9195/2020\|EPI_ISL_570230\|2020-05 | .................... | .................... | .................... | .................... | .................... | .................... |
| K | 1249 | hCoV-19/USA/WA-UW-9210/2020\|EPI_ISL_570231\|2020-05 | .................... | .................... | .................... | .................... | .................... | .................... |
| K | 1250 | hCoV-19/USA/WA-UW-9214/2020\|EPI_ISL_570232\|2020-05 | .................... | .................... | .................... | .................... | .................... | .................... |
| K | 1251 | hCoV-19/USA/WA-UW-9227/2020\|EPI_ISL_570233\|2020-05 | .................... | .................... | .................... | .................... | .................... | .................... |
| K | 1252 | hCoV-19/USA/WA-UW-9228/2020\|EPI_ISL_570234\|2020-05 | .................... | .................... | .................... | .................... | .................... | .................... |
| K | 1253 | hCoV-19/USA/WA-UW-9229/2020\|EPI_ISL_570235\|2020-05 | .................... | .................... | .................... | .................... | .................... | .................... |
| K | 1254 | hCoV-19/USA/WA-UW-9230/2020\|EPI_ISL_570236\|2020-05 | .................... | .................... | .................... | .................... | .................... | .................... |
| K | 1255 | hCoV-19/USA/WA-UW-9233/2020\|EPI_ISL_570237\|2020-05 | .................... | .................... | .................... | .................... | .................... | .................... |
| K | 1256 | hCoV-19/USA/WA-UW-9235/2020\|EPI_ISL_570238\|2020-05 | .................... | .................... | .................... | .................... | .................... | .................... |
| K | 1257 | hCoV-19/USA/WA-UW-9244/2020\|EPI_ISL_570239\|2020-05 | .................... | .................... | .................... | .................... | .................... | .................... |
| K | 1258 | hCoV-19/USA/WA-UW-9255/2020\|EPI_ISL_570240\|2020-05 | .................... | .................... | .................... | .................... | .................... | .................... |
| K | 1259 | hCoV-19/USA/WA-UW-9266/2020\|EPI_ISL_570241\|2020-05 | .................... | .................... | .................... | .................... | .................... | .................... |
| K | 1260 | hCoV-19/USA/WA-UW-10960/2020\|EPI_ISL_570363\|2020-0 | .................... | .................... | .................... | .................... | .................... | .................... |

| | | | | | | | |
|---|---|---|---|---|---|---|---|
| K | 1261 hCoV-19/USA/WA-UW-11002/2020\|EPI_ISL_570364\|2020-0 | .................... | .................... | .................... | .................... | .................... | .................... |
| K | 1262 hCoV-19/USA/WA-UW-11038/2020\|EPI_ISL_570366\|2020-0 | .................... | .................... | .................... | .................... | .................... | .................... |
| K | 1263 hCoV-19/USA/WA-UW-11040/2020\|EPI_ISL_570367\|2020-0 | .................... | .................... | .................... | .................... | .................... | .................... |
| K | 1264 hCoV-19/USA/WA-UW-11043/2020\|EPI_ISL_570368\|2020-0 | .................... | .................... | .................... | .................... | .................... | .................... |
| K | 1265 hCoV-19/USA/WA-UW-11059/2020\|EPI_ISL_570369\|2020-0 | .................... | .................... | .................... | .................... | .................... | .................... |
| K | 1266 hCoV-19/USA/WA-UW-11278/2020\|EPI_ISL_570385\|2020-0 | .................... | .................... | .................... | .................... | .................... | .................... |
| K | 1267 hCoV-19/USA/WA-UW-11280/2020\|EPI_ISL_570386\|2020-0 | .................... | .................... | .................... | .................... | .................... | .................... |
| K | 1268 hCoV-19/USA/WA-UW-11285/2020\|EPI_ISL_570387\|2020-0 | .................... | .................... | .................... | .................... | .................... | .................... |
| K | 1269 hCoV-19/USA/WA-UW-11287/2020\|EPI_ISL_570388\|2020-0 | .................... | .................... | .................... | .................... | .................... | .................... |
| K | 1270 hCoV-19/USA/WA-UW-11289/2020\|EPI_ISL_570389\|2020-0 | .................... | .................... | .................... | .................... | .....T.............. | .................... |
| K | 1271 hCoV-19/USA/WA-UW-11290/2020\|EPI_ISL_570390\|2020-0 | .................... | .................... | .................... | .................... | .................... | .................... |
| K | 1272 hCoV-19/USA/WA-UW-11297/2020\|EPI_ISL_570391\|2020-0 | .................... | .................... | .................... | .................... | .................... | .................... |
| K | 1273 hCoV-19/USA/WA-UW-11309/2020\|EPI_ISL_570392\|2020-0 | .................... | .................... | .................... | .................... | .................... | .................... |
| K | 1274 hCoV-19/USA/WA-UW-11324/2020\|EPI_ISL_570393\|2020-0 | .................... | .A.................. | .................... | .................... | .................... | .................... |
| K | 1275 hCoV-19/USA/WA-UW-11325/2020\|EPI_ISL_570394\|2020-0 | .................... | .A.................. | .................... | .................... | .................... | .................... |
| K | 1276 hCoV-19/USA/WA-UW-11328/2020\|EPI_ISL_570395\|2020-0 | .................... | .................... | .................... | .................... | .................... | .................... |
| K | 1277 hCoV-19/USA/WA-UW-11338/2020\|EPI_ISL_570396\|2020-0 | .................... | .................... | .................... | .................... | .................... | .................... |
| K | 1278 hCoV-19/USA/WA-UW-11341/2020\|EPI_ISL_570397\|2020-0 | .................... | .................... | .................... | .................... | .................... | .................... |
| K | 1279 hCoV-19/USA/WA-UW-11347/2020\|EPI_ISL_570398\|2020-0 | .................... | .................... | .................... | .................... | .................... | .................... |
| K | 1280 hCoV-19/USA/WA-UW-11349/2020\|EPI_ISL_570399\|2020-0 | .................... | .................... | .................... | .................... | .................... | .................... |
| K | 1281 hCoV-19/USA/MN-MDH-1713/2020\|EPI_ISL_560800\|2020-0 | .................... | .................... | .................... | .................... | .................... | .................... |
| K | 1282 hCoV-19/USA/MN-MDH-1714/2020\|EPI_ISL_560801\|2020-0 | .................... | .................... | .................... | .................... | .................... | .................... |
| K | 1283 hCoV-19/USA/MN-MDH-1715/2020\|EPI_ISL_560802\|2020-0 | .................... | .................... | .................... | .................... | .................... | .................... |
| K | 1284 hCoV-19/USA/MN-MDH-1716/2020\|EPI_ISL_560803\|2020-0 | .................... | .................... | .................... | .................... | .................... | .................... |
| K | 1285 hCoV-19/USA/MN-MDH-1717/2020\|EPI_ISL_560804\|2020-0 | .................... | .A.................. | .................... | .................... | .................... | .................... |
| K | 1286 hCoV-19/USA/MN-MDH-1718/2020\|EPI_ISL_560805\|2020-0 | .................... | .A.................. | .................... | .................... | .................... | .................... |
| K | 1287 hCoV-19/USA/MD-MDH-0193/2020\|EPI_ISL_560806\|2020-0 | .................... | .................... | .................... | .................... | .................... | .................... |
| K | 1288 hCoV-19/USA/MD-MDH-0194/2020\|EPI_ISL_560807\|2020-0 | .................... | .................... | .................... | .................... | .................... | .................... |
| K | 1289 hCoV-19/USA/MD-MDH-0195/2020\|EPI_ISL_560808\|2020-0 | .................... | .................... | .................... | .................... | .................... | .................... |
| K | 1290 hCoV-19/USA/MD-MDH-0196/2020\|EPI_ISL_560809\|2020-0 | .................... | .................... | .................... | .................... | .................... | .................... |
| K | 1291 hCoV-19/USA/MD-MDH-0197/2020\|EPI_ISL_560810\|2020-0 | .................... | .................... | .................... | .................... | .................... | .................... |
| K | 1292 hCoV-19/USA/MD-MDH-0198/2020\|EPI_ISL_560811\|2020-0 | .................... | .................... | .................... | .................... | .................... | .................... |
| K | 1293 hCoV-19/USA/MD-MDH-0199/2020\|EPI_ISL_560812\|2020-0 | .................... | .................... | .................... | .................... | .................... | .................... |
| K | 1294 hCoV-19/USA/MD-MDH-0200/2020\|EPI_ISL_560813\|2020-0 | .................... | .................... | .................... | .................... | .................... | .................... |
| K | 1295 hCoV-19/USA/MD-MDH-0201/2020\|EPI_ISL_560814\|2020-0 | .................... | .................... | .................... | .................... | .................... | .................... |
| K | 1296 hCoV-19/USA/MD-MDH-0202/2020\|EPI_ISL_560815\|2020-0 | .................... | .................... | .................... | .................... | .................... | .................... |
| K | 1297 hCoV-19/USA/MD-MDH-0203/2020\|EPI_ISL_560816\|2020-0 | .................... | .................... | .................... | .................... | .................... | .................... |
| K | 1298 hCoV-19/USA/MD-MDH-0204/2020\|EPI_ISL_560817\|2020-0 | .................... | .................... | .................... | .................... | .................... | .................... |
| K | 1299 hCoV-19/USA/MD-MDH-0205/2020\|EPI_ISL_560818\|2020-0 | .................... | .................... | .................... | .................... | .................... | .................... |
| K | 1300 hCoV-19/USA/MD-MDH-0206/2020\|EPI_ISL_560819\|2020-0 | .................... | .................... | .................... | .................... | .................... | .................... |
| K | 1301 hCoV-19/USA/MD-MDH-0207/2020\|EPI_ISL_560820\|2020-0 | .................... | .................... | .................... | .................... | .................... | .................... |
| K | 1302 hCoV-19/USA/MD-MDH-0208/2020\|EPI_ISL_560821\|2020-0 | .................... | .................... | .................... | .................... | .................... | .................... |
| K | 1303 hCoV-19/USA/MD-MDH-0209/2020\|EPI_ISL_560822\|2020-0 | .................... | .................... | .................... | .................... | .................... | .................... |
| K | 1304 hCoV-19/USA/MD-MDH-0210/2020\|EPI_ISL_560823\|2020-0 | .................... | .................... | .................... | .................... | .................... | .................... |
| K | 1305 hCoV-19/USA/MD-MDH-0211/2020\|EPI_ISL_560824\|2020-0 | .................... | .................... | .................... | .................... | .................... | .................... |
| K | 1306 hCoV-19/USA/MD-MDH-0212/2020\|EPI_ISL_560825\|2020-0 | .................... | .................... | .................... | .................... | .................... | .................... |
| K | 1307 hCoV-19/USA/MD-MDH-0213/2020\|EPI_ISL_560826\|2020-0 | .................... | .................... | .................... | .................... | .................... | .................... |
| K | 1308 hCoV-19/USA/MD-MDH-0214/2020\|EPI_ISL_560827\|2020-0 | .................... | .................... | .................... | .................... | .................... | .................... |
| K | 1309 hCoV-19/USA/MD-MDH-0215/2020\|EPI_ISL_560828\|2020-0 | .................... | .................... | .................... | .................... | .................... | .................... |
| K | 1310 hCoV-19/USA/MD-MDH-0216/2020\|EPI_ISL_560829\|2020-0 | .................... | .................... | .................... | .................... | .................... | .................... |
| K | 1311 hCoV-19/USA/MD-MDH-0217/2020\|EPI_ISL_560830\|2020-0 | .................... | .A.................. | .................... | .................... | .................... | .................... |
| K | 1312 hCoV-19/USA/MD-MDH-0218/2020\|EPI_ISL_560831\|2020-0 | .................... | .................... | .................... | .................... | .................... | .................... |
| K | 1313 hCoV-19/USA/MD-MDH-0219/2020\|EPI_ISL_560832\|2020-0 | .................... | .................... | .................... | .................... | .................... | .................... |
| K | 1314 hCoV-19/USA/UT-UPHL-04427/2020\|EPI_ISL_560837\|2020 | .................... | .................... | .................... | .................... | .................... | .................... |
| K | 1315 hCoV-19/USA/UT-UPHL-04426/2020\|EPI_ISL_560838\|2020 | .................... | .................... | .................... | .................... | .................... | .................... |
| K | 1316 hCoV-19/Australia/VIC10696/2020\|EPI_ISL_563037\|202 | .................... | .................... | .................... | .................... | .................... | .................... |
| K | 1317 hCoV-19/Australia/VIC10698/2020\|EPI_ISL_563038\|202 | .................... | .................... | .................... | .................... | .................... | .................... |
| K | 1318 hCoV-19/Australia/VIC10700/2020\|EPI_ISL_563039\|202 | .................... | .................... | .................... | .................... | .................... | .................... |
| K | 1319 hCoV-19/Australia/VIC10701/2020\|EPI_ISL_563040\|202 | .................... | .................... | .................... | .................... | .................... | .................... |
| K | 1320 hCoV-19/Australia/VIC10702/2020\|EPI_ISL_563041\|202 | .................... | .................... | .................... | .................... | .................... | .................... |
| K | 1321 hCoV-19/Australia/VIC10703/2020\|EPI_ISL_563042\|202 | .................... | .................... | .................... | .................... | .................... | .................... |
| K | 1322 hCoV-19/Australia/VIC10704/2020\|EPI_ISL_563043\|202 | .................... | .................... | .................... | .................... | .................... | ...............A.... |
| K | 1323 hCoV-19/Australia/VIC10705/2020\|EPI_ISL_563044\|202 | .................... | .................... | .................... | .................... | .................... | .................... |
| K | 1324 hCoV-19/Australia/VIC10706/2020\|EPI_ISL_563045\|202 | .................... | .................... | .................... | .................... | .................... | .................... |
| K | 1325 hCoV-19/Australia/VIC10708/2020\|EPI_ISL_563046\|202 | .................... | .................... | .................... | .................... | .................... | .................... |
| K | 1326 hCoV-19/Australia/VIC10709/2020\|EPI_ISL_563047\|202 | .................... | .................... | .................... | .................... | .................... | .................... |
| K | 1327 hCoV-19/Australia/VIC10716/2020\|EPI_ISL_563050\|202 | .................... | .................... | .................... | .................... | .................... | .................... |
| K | 1328 hCoV-19/Australia/VIC10731/2020\|EPI_ISL_563059\|202 | .................... | .................... | .................... | .................... | .................... | .................... |
| K | 1329 hCoV-19/Australia/VIC10736/2020\|EPI_ISL_563060\|202 | .................... | .................... | .................... | .................... | .................... | .................... |
| K | 1330 hCoV-19/Australia/VIC10737/2020\|EPI_ISL_563061\|202 | .................... | .................... | .................... | .................... | .................... | .................... |
| K | 1331 hCoV-19/Australia/VIC10738/2020\|EPI_ISL_563062\|202 | .................... | .................... | .................... | .................... | .................... | .................... |
| K | 1332 hCoV-19/Australia/VIC10740/2020\|EPI_ISL_563063\|202 | .................... | .................... | .................... | .................... | .................... | .................... |
| K | 1333 hCoV-19/Australia/VIC10742/2020\|EPI_ISL_563064\|202 | .................... | .................... | .................... | .................... | .................... | .................... |
| K | 1334 hCoV-19/Australia/VIC10743/2020\|EPI_ISL_563065\|202 | .................... | .................... | .................... | .................... | .................... | .................... |
| K | 1336 hCoV-19/Australia/VIC10747/2020\|EPI_ISL_563067\|202 | .................... | .................... | .................... | .................... | .................... | .................... |
| K | 1337 hCoV-19/Australia/VIC10750/2020\|EPI_ISL_563068\|202 | .................... | .................... | .................... | .................... | .................... | .................... |
| K | 1338 hCoV-19/Australia/VIC10751/2020\|EPI_ISL_563069\|202 | .................... | .................... | .................... | .................... | .................... | .................... |
| K | 1339 hCoV-19/Australia/VIC10752/2020\|EPI_ISL_563070\|202 | .................... | .................... | .................... | .................... | .................... | .................... |
| K | 1340 hCoV-19/Australia/VIC10753/2020\|EPI_ISL_563071\|202 | .................... | .................... | .................... | .................... | .................... | .................... |
| K | 1342 hCoV-19/Australia/VIC10756/2020\|EPI_ISL_563073\|202 | .................... | .................... | .................... | .................... | .................... | .................... |

| | | | | | | | | | |
|---|---|---|---|---|---|---|---|---|---|
| K | 1343 | hCoV-19/Australia/VIC10757/2020\|EPI_ISL_563074\|202 | ···················· | ···················· | ···················· | ···················· | ···················· | ···················· | |
| K | 1344 | hCoV-19/Australia/VIC10758/2020\|EPI_ISL_563075\|202 | ···················· | ···················· | ···················· | ···················· | ···················· | ···················· | |
| K | 1345 | hCoV-19/Australia/VIC10759/2020\|EPI_ISL_563076\|202 | ···················· | ···················· | ···················· | ···················· | ···················· | ···················· | |
| K | 1346 | hCoV-19/Australia/VIC10760/2020\|EPI_ISL_563077\|202 | ···················· | ···················· | ···················· | ···················· | ···················· | ···················· | |
| K | 1347 | hCoV-19/Australia/VIC10761/2020\|EPI_ISL_563078\|202 | ···················· | ···················· | ···················· | ···················· | ···················· | ···················· | |
| K | 1348 | hCoV-19/Australia/VIC10762/2020\|EPI_ISL_563079\|202 | ···················· | ···················· | ···················· | ···················· | ···················· | ···················· | |
| K | 1349 | hCoV-19/Australia/VIC10770/2020\|EPI_ISL_563080\|202 | ···················· | ···················· | ···················· | ···················· | ···················· | ···················· | |
| K | 1350 | hCoV-19/Australia/VIC10771/2020\|EPI_ISL_563081\|202 | ···················· | ···················· | ···················· | ···················· | ···················· | ···················· | |
| K | 1351 | hCoV-19/Australia/VIC10773/2020\|EPI_ISL_563082\|202 | ···················· | ···················· | ···················· | ···················· | ···················· | ···················· | |
| K | 1352 | hCoV-19/Australia/VIC10774/2020\|EPI_ISL_563083\|202 | ···················· | ···················· | ···················· | ···················· | ···················· | ···················· | |
| K | 1353 | hCoV-19/Australia/VIC10775/2020\|EPI_ISL_563084\|202 | ···················· | ···················· | ···················· | ···················· | ···················· | ···················· | |
| K | 1354 | hCoV-19/Australia/VIC10777/2020\|EPI_ISL_563085\|202 | ···················· | ···················· | ···················· | ···················· | ···················· | ···················· | |
| K | 1355 | hCoV-19/Australia/VIC10781/2020\|EPI_ISL_563086\|202 | ···················· | ···················· | ···················· | ···················· | ···················· | ···················· | |
| K | 1356 | hCoV-19/Australia/VIC10782/2020\|EPI_ISL_563087\|202 | ···················· | ···················· | ···················· | ···················· | ···················· | ···················· | |
| K | 1357 | hCoV-19/Australia/VIC10786/2020\|EPI_ISL_563088\|202 | ···················· | ···················· | ···················· | ···················· | ···················· | ···················· | |
| K | 1359 | hCoV-19/Australia/VIC10788/2020\|EPI_ISL_563090\|202 | ···················· | ···················· | ···················· | ···················· | ···················· | ···················· | |
| K | 1361 | hCoV-19/Australia/VIC10792/2020\|EPI_ISL_563092\|202 | ···················· | ···················· | ···················· | ···················· | ···················· | ···················· | |
| K | 1362 | hCoV-19/Australia/VIC10794/2020\|EPI_ISL_563093\|202 | ···················· | ···················· | ···················· | ···················· | ···················· | ···················· | |
| K | 1363 | hCoV-19/Australia/VIC10795/2020\|EPI_ISL_563094\|202 | ···················· | ···················· | ···················· | ···················· | ···················· | ···················· | |
| K | 1364 | hCoV-19/Australia/VIC10796/2020\|EPI_ISL_563095\|202 | ···················· | ···················· | ···················· | ···················· | ···················· | ···················· | |
| K | 1365 | hCoV-19/Australia/VIC10798/2020\|EPI_ISL_563096\|202 | ···················· | ···················· | ···················· | ···················· | ···················· | ···················· | |
| K | 1366 | hCoV-19/Australia/VIC10800/2020\|EPI_ISL_563097\|202 | ···················· | ···················· | ···················· | ···················· | ···················· | ···················· | |
| K | 1367 | hCoV-19/Australia/VIC10801/2020\|EPI_ISL_563098\|202 | ···················· | ···················· | ···················· | ···················· | ···················· | ···················· | |
| K | 1368 | hCoV-19/Australia/VIC10806/2020\|EPI_ISL_563099\|202 | ···················· | ···················· | ···················· | ···················· | ···················· | ···················· | |
| K | 1369 | hCoV-19/Australia/VIC10808/2020\|EPI_ISL_563100\|202 | ···················· | ···················· | ···················· | ···················· | ···················· | ···················· | |
| K | 1370 | hCoV-19/Australia/VIC10809/2020\|EPI_ISL_563101\|202 | ···················· | ···················· | ···················· | ···················· | ···················· | ···················· | |
| K | 1371 | hCoV-19/Australia/VIC10811/2020\|EPI_ISL_563102\|202 | ···················· | ···················· | ···················· | ···················· | ···················· | ···················· | |
| K | 1372 | hCoV-19/Australia/VIC10812/2020\|EPI_ISL_563103\|202 | ···················· | ···················· | ···················· | ···················· | ···················· | ···················· | |
| K | 1373 | hCoV-19/Australia/VIC10813/2020\|EPI_ISL_563104\|202 | ···················· | ···················· | ···················· | ···················· | ···················· | ···················· | |
| K | 1374 | hCoV-19/Australia/VIC10814/2020\|EPI_ISL_563105\|202 | ···················· | ···················· | ···················· | ···················· | ···················· | ···················· | |
| K | 1375 | hCoV-19/Australia/VIC10817/2020\|EPI_ISL_563106\|202 | ···················· | ···················· | ···················· | ···················· | ···················· | ···················· | |
| K | 1377 | hCoV-19/Australia/VIC10819/2020\|EPI_ISL_563108\|202 | ···················· | ···················· | ···················· | ···················· | ···················· | ···················· | |
| K | 1378 | hCoV-19/Australia/VIC10820/2020\|EPI_ISL_563109\|202 | ···················· | ···················· | ···················· | ···················· | ···················· | ···················· | |
| K | 1380 | hCoV-19/Australia/VIC10822/2020\|EPI_ISL_563111\|202 | ···················· | ···················· | ···················· | ···················· | ···················· | ···················· | |
| K | 1381 | hCoV-19/Australia/VIC10825/2020\|EPI_ISL_563112\|202 | ···················· | ···················· | ···················· | ···················· | ···················· | ···················· | |
| K | 1382 | hCoV-19/Australia/VIC10828/2020\|EPI_ISL_563113\|202 | ···················· | ···················· | ···················· | ···················· | ···················· | ···················· | |
| K | 1383 | hCoV-19/Australia/VIC10829/2020\|EPI_ISL_563114\|202 | ···················· | ···················· | ···················· | ···················· | ···················· | ···················· | |
| K | 1384 | hCoV-19/Australia/VIC10830/2020\|EPI_ISL_563115\|202 | ···················· | ···················· | ···················· | ···················· | ···················· | ·NNN················ | 1 |
| K | 1385 | hCoV-19/Australia/VIC10833/2020\|EPI_ISL_563118\|202 | ···················· | ···················· | ···················· | ···················· | ···················· | ···················· | |
| K | 1386 | hCoV-19/Australia/VIC10835/2020\|EPI_ISL_563119\|202 | ···················· | ···················· | ···················· | ···················· | ···················· | ···················· | |
| K | 1388 | hCoV-19/Australia/VIC10849/2020\|EPI_ISL_563126\|202 | ···················· | ···················· | ···················· | ···················· | ···················· | ···················· | |
| K | 1390 | hCoV-19/Australia/VIC10857/2020\|EPI_ISL_563128\|202 | ···················· | ···················· | ···················· | ···················· | ···················· | ···················· | |
| K | 1391 | hCoV-19/Australia/VIC10858/2020\|EPI_ISL_563129\|202 | ···················· | ···················· | ···················· | ···················· | ···················· | ···················· | |
| K | 1392 | hCoV-19/Australia/VIC10863/2020\|EPI_ISL_563130\|202 | ···················· | ···················· | ···················· | ···················· | ···················· | ···················· | |
| K | 1393 | hCoV-19/Australia/VIC10866/2020\|EPI_ISL_563131\|202 | ···················· | ···················· | ···················· | ···················· | ···················· | ···················· | |
| K | 1394 | hCoV-19/Australia/VIC10867/2020\|EPI_ISL_563132\|202 | ···················· | ···················· | ···················· | ···················· | ···················· | ···················· | |
| K | 1395 | hCoV-19/Australia/VIC10869/2020\|EPI_ISL_563133\|202 | ···················· | ···················· | ···················· | ···················· | ···················· | ···················· | |
| K | 1396 | hCoV-19/Australia/VIC10871/2020\|EPI_ISL_563134\|202 | ···················· | ···················· | ···················· | ···················· | ···················· | ···················· | |
| K | 1397 | hCoV-19/Australia/VIC10874/2020\|EPI_ISL_563135\|202 | ···················· | ···················· | ···················· | ···················· | ···················· | ···················· | |
| K | 1398 | hCoV-19/Australia/VIC7378/2020\|EPI_ISL_564368\|2020 | ···················· | ···················· | ···················· | ···················· | ···················· | ···················· | |
| K | 1399 | hCoV-19/Australia/VIC7380/2020\|EPI_ISL_564369\|2020 | ···················· | ···················· | ···················· | ···················· | ···················· | ···················· | |
| K | 1400 | hCoV-19/Australia/VIC7381/2020\|EPI_ISL_564370\|2020 | ···················· | ···················· | ···················· | ···················· | ···················· | ···················· | |
| K | 1401 | hCoV-19/Australia/VIC7382/2020\|EPI_ISL_564371\|2020 | ···················· | ···················· | ···················· | ···················· | ···················· | ···················· | |
| K | 1402 | hCoV-19/Australia/VIC7383/2020\|EPI_ISL_564372\|2020 | ···················· | ···················· | ···················· | ···················· | ···················· | ···················· | |
| K | 1403 | hCoV-19/Australia/VIC7386/2020\|EPI_ISL_564373\|2020 | ···················· | ···················· | ···················· | ···················· | ···················· | ···················· | |
| K | 1404 | hCoV-19/Australia/VIC7388/2020\|EPI_ISL_564374\|2020 | ···················· | ···················· | ···················· | ···················· | ···················· | ···················· | |
| K | 1405 | hCoV-19/Australia/VIC7389/2020\|EPI_ISL_564375\|2020 | ···················· | ···················· | ···················· | ···················· | ···················· | ···················· | |
| K | 1406 | hCoV-19/Australia/VIC7390/2020\|EPI_ISL_564376\|2020 | ···················· | ···················· | ···················· | ···················· | ···················· | ···················· | |
| K | 1407 | hCoV-19/Australia/VIC7391/2020\|EPI_ISL_564377\|2020 | ···················· | ···················· | ···················· | ···················· | ···················· | ···················· | |
| K | 1408 | hCoV-19/Australia/VIC7392/2020\|EPI_ISL_564378\|2020 | ···················· | ···················· | ···················· | ···················· | ···················· | ···················· | |
| K | 1409 | hCoV-19/Australia/VIC7411/2020\|EPI_ISL_564390\|2020 | ···················· | ···················· | ···················· | ···················· | ···················· | ···················· | |
| K | 1410 | hCoV-19/Australia/VIC7414/2020\|EPI_ISL_564391\|2020 | ···················· | ···················· | ···················· | ···················· | ···················· | ···················· | |
| K | 1411 | hCoV-19/Australia/VIC7415/2020\|EPI_ISL_564392\|2020 | ···················· | ···················· | ···················· | ···················· | ···················· | ···················· | |
| K | 1412 | hCoV-19/Australia/VIC7416/2020\|EPI_ISL_564393\|2020 | ···················· | ···················· | ···················· | ···················· | ···················· | ···················· | |
| K | 1413 | hCoV-19/Australia/VIC7417/2020\|EPI_ISL_564394\|2020 | ···················· | ···················· | ···················· | ···················· | ···················· | ···················· | |
| K | 1414 | hCoV-19/Australia/VIC7418/2020\|EPI_ISL_564395\|2020 | ···················· | ···················· | ···················· | ···················· | ···················· | ···················· | |
| K | 1415 | hCoV-19/Australia/VIC7419/2020\|EPI_ISL_564396\|2020 | ···················· | ···················· | ···················· | ···················· | ···················· | ···················· | |
| K | 1416 | hCoV-19/Australia/VIC7420/2020\|EPI_ISL_564397\|2020 | ···················· | ···················· | ···················· | ···················· | ···················· | ···················· | |
| K | 1417 | hCoV-19/Australia/VIC7421/2020\|EPI_ISL_564398\|2020 | ···················· | ···················· | ···················· | ···················· | ···················· | ···················· | |
| K | 1418 | hCoV-19/Australia/VIC7423/2020\|EPI_ISL_564399\|2020 | ···················· | ···················· | ···················· | ···················· | ···················· | ···················· | |
| K | 1419 | hCoV-19/Australia/VIC7425/2020\|EPI_ISL_564401\|2020 | ···················· | ···················· | ···················· | ···················· | ···················· | ···················· | |
| K | 1420 | hCoV-19/Australia/VIC7426/2020\|EPI_ISL_564402\|2020 | ···················· | ···················· | ···················· | ···················· | ···················· | ···················· | |
| K | 1421 | hCoV-19/Australia/VIC7427/2020\|EPI_ISL_564403\|2020 | ···················· | ···················· | ···················· | ···················· | ···················· | ···················· | |
| K | 1422 | hCoV-19/Australia/VIC7428/2020\|EPI_ISL_564404\|2020 | ···················· | ···················· | ···················· | ···················· | ···················· | ···················· | |
| K | 1423 | hCoV-19/Australia/VIC7429/2020\|EPI_ISL_564405\|2020 | ···················· | ···················· | ···················· | ···················· | ···················· | ···················· | |
| K | 1424 | hCoV-19/Australia/VIC7430/2020\|EPI_ISL_564406\|2020 | ···················· | ···················· | ···················· | ···················· | ···················· | ···················· | |
| K | 1425 | hCoV-19/Australia/VIC7431/2020\|EPI_ISL_564407\|2020 | ···················· | ···················· | ···················· | ···················· | ···················· | ···················· | |
| K | 1426 | hCoV-19/Australia/VIC7432/2020\|EPI_ISL_564408\|2020 | ···················· | ···················· | ···················· | ···················· | ···················· | ···················· | |
| K | 1427 | hCoV-19/Australia/VIC7433/2020\|EPI_ISL_564409\|2020 | ···················· | ···················· | ···················· | ···················· | ···················· | ···················· | |
| K | 1428 | hCoV-19/Australia/VIC7434/2020\|EPI_ISL_564410\|2020 | ···················· | ···················· | ···················· | ···················· | ···················· | ···················· | |

| | | | | | | | | |
|---|---|---|---|---|---|---|---|---|
| K | 1429 | hCoV-19/Australia/VIC7435/2020\|EPI_ISL_564411\|2020 | ···················· | ···················· | ····················· | ···················· | ···················· | ···················· |
| K | 1430 | hCoV-19/Australia/VIC7436/2020\|EPI_ISL_564412\|2020 | ···················· | ···················· | ····················· | ···················· | ···················· | ···················· |
| K | 1431 | hCoV-19/Australia/VIC7438/2020\|EPI_ISL_564413\|2020 | ···················· | ···················· | ····················· | ···················· | ···················· | ···················· |
| K | 1432 | hCoV-19/Australia/VIC7439/2020\|EPI_ISL_564414\|2020 | ···················· | ···················· | ····················· | ···················· | ···················· | ···················· |
| K | 1433 | hCoV-19/Australia/VIC7441/2020\|EPI_ISL_564415\|2020 | ···················· | ···················· | ····················· | ···················· | ···················· | ···················· |
| K | 1434 | hCoV-19/Australia/VIC7442/2020\|EPI_ISL_564416\|2020 | ···················· | ···················· | ····················· | ···················· | ···················· | ···················· |
| K | 1435 | hCoV-19/Australia/VIC7443/2020\|EPI_ISL_564417\|2020 | ···················· | ···················· | ····················· | ···················· | ···················· | ···················· |
| K | 1436 | hCoV-19/Australia/VIC7444/2020\|EPI_ISL_564418\|2020 | ···················· | ···················· | ····················· | ···················· | ···················· | ···················· |
| K | 1437 | hCoV-19/Australia/VIC7445/2020\|EPI_ISL_564419\|2020 | ···················· | ···················· | ····················· | ···················· | ···················· | ···················· |
| K | 1438 | hCoV-19/Australia/VIC7448/2020\|EPI_ISL_564420\|2020 | ···················· | ···················· | ····················· | ···················· | ···················· | ···················· |
| K | 1439 | hCoV-19/Australia/VIC7449/2020\|EPI_ISL_564421\|2020 | ···················· | ···················· | ····················· | ···················· | ···················· | ···················· |
| K | 1441 | hCoV-19/Australia/VIC7452/2020\|EPI_ISL_564423\|2020 | ···················· | ···················· | ····················· | ···················· | ···················· | ···················· |
| K | 1442 | hCoV-19/Australia/VIC7453/2020\|EPI_ISL_564424\|2020 | ···················· | ···················· | ····················· | ···················· | ···················· | ···················· |
| K | 1443 | hCoV-19/Australia/VIC7454/2020\|EPI_ISL_564425\|2020 | ···················· | ···················· | ····················· | ···················· | ···················· | ···················· |
| K | 1444 | hCoV-19/Australia/VIC7455/2020\|EPI_ISL_564426\|2020 | ···················· | ···················· | ····················· | ···················· | ···················· | ···················· |
| K | 1445 | hCoV-19/Australia/VIC7456/2020\|EPI_ISL_564427\|2020 | ···················· | ···················· | ····················· | ···················· | ···················· | ···················· |
| K | 1446 | hCoV-19/Australia/VIC7457/2020\|EPI_ISL_564428\|2020 | ···················· | ···················· | ····················· | ···················· | ···················· | ···················· |
| K | 1447 | hCoV-19/Australia/VIC7459/2020\|EPI_ISL_564429\|2020 | ···················· | ···················· | ····················· | ···················· | ···················· | ···················· |
| K | 1448 | hCoV-19/Australia/VIC7467/2020\|EPI_ISL_564430\|2020 | ···················· | ···················· | ····················· | ···················· | ···················· | ···················· |
| K | 1449 | hCoV-19/Australia/VIC7471/2020\|EPI_ISL_564431\|2020 | ···················· | ···················· | ····················· | ···················· | ···················· | ···················· |
| K | 1450 | hCoV-19/Australia/VIC7472/2020\|EPI_ISL_564432\|2020 | ···················· | ···················· | ····················· | ···················· | ···················· | ···················· |
| K | 1451 | hCoV-19/Australia/VIC7473/2020\|EPI_ISL_564433\|2020 | ···················· | ···················· | ····················· | ···················· | ···················· | ···················· |
| K | 1452 | hCoV-19/Australia/VIC7475/2020\|EPI_ISL_564434\|2020 | ···················· | ···················· | ····················· | ···················· | ···················· | ···················· |
| K | 1453 | hCoV-19/Australia/VIC7476/2020\|EPI_ISL_564435\|2020 | ···················· | ···················· | ····················· | ···················· | ···················· | ···················· |
| K | 1454 | hCoV-19/Australia/VIC7477/2020\|EPI_ISL_564436\|2020 | ···················· | ···················· | ····················· | ···················· | ···················· | ···················· |
| K | 1455 | hCoV-19/Australia/VIC7478/2020\|EPI_ISL_564437\|2020 | ···················· | ···················· | ····················· | ···················· | ···················· | ···················· |
| K | 1456 | hCoV-19/Australia/VIC7481/2020\|EPI_ISL_564438\|2020 | ···················· | ···················· | ····················· | ···················· | ···················· | ···················· |
| K | 1457 | hCoV-19/Australia/VIC7482/2020\|EPI_ISL_564439\|2020 | ···················· | ···················· | ····················· | ···················· | ···················· | ···················· |
| K | 1458 | hCoV-19/Australia/VIC7484/2020\|EPI_ISL_564440\|2020 | ···················· | ···················· | ····················· | ···················· | ···················· | ···················· |
| K | 1459 | hCoV-19/Australia/VIC7485/2020\|EPI_ISL_564441\|2020 | ···················· | ···················· | ····················· | ···················· | ···················· | ···················· |
| K | 1460 | hCoV-19/Australia/VIC7486/2020\|EPI_ISL_564442\|2020 | ···················· | ···················· | ····················· | ···················· | ···················· | ···················· |
| K | 1461 | hCoV-19/Australia/VIC7489/2020\|EPI_ISL_564443\|2020 | ···················· | ···················· | ····················· | ···················· | ···················· | ···················· |
| K | 1462 | hCoV-19/Australia/VIC7491/2020\|EPI_ISL_564444\|2020 | ···················· | ···················· | ····················· | ···················· | ···················· | ···················· |
| K | 1463 | hCoV-19/Australia/VIC7492/2020\|EPI_ISL_564445\|2020 | ···················· | ···················· | ····················· | ···················· | ···················· | ···················· |
| K | 1464 | hCoV-19/Australia/VIC7493/2020\|EPI_ISL_564446\|2020 | ···················· | ···················· | ····················· | ···················· | ···················· | ···················· |
| K | 1465 | hCoV-19/Australia/VIC7494/2020\|EPI_ISL_564447\|2020 | ···················· | ···················· | ····················· | ···················· | ···················· | ···················· |
| K | 1466 | hCoV-19/Australia/VIC7496/2020\|EPI_ISL_564449\|2020 | ···················· | ···················· | ····················· | ···················· | ···················· | ···················· |
| K | 1467 | hCoV-19/Australia/VIC7508/2020\|EPI_ISL_564456\|2020 | ···················· | ···················· | ····················· | ···················· | ···················· | ···················· |
| K | 1468 | hCoV-19/Australia/VIC7509/2020\|EPI_ISL_564457\|2020 | ···················· | ···················· | ····················· | ···················· | ···················· | ···················· |
| K | 1469 | hCoV-19/Australia/VIC7511/2020\|EPI_ISL_564458\|2020 | ···················· | ···················· | ····················· | ···················· | ···················· | ···················· |
| K | 1470 | hCoV-19/Australia/VIC7512/2020\|EPI_ISL_564459\|2020 | ···················· | ···················· | ····················· | ···················· | ···················· | ···················· |
| K | 1471 | hCoV-19/Australia/VIC7513/2020\|EPI_ISL_564460\|2020 | ···················· | ···················· | ····················· | ···················· | ···················· | ···················· |
| K | 1472 | hCoV-19/Australia/VIC7514/2020\|EPI_ISL_564461\|2020 | ···················· | ···················· | ····················· | ···················· | ···················· | ···················· |
| K | 1473 | hCoV-19/Australia/VIC7515/2020\|EPI_ISL_564462\|2020 | ···················· | ···················· | ····················· | ···················· | ···················· | ···················· |
| K | 1474 | hCoV-19/Australia/VIC7517/2020\|EPI_ISL_564463\|2020 | ···················· | ···················· | ····················· | ···················· | ···················· | ···················· |
| K | 1475 | hCoV-19/Australia/VIC7518/2020\|EPI_ISL_564464\|2020 | ···················· | ···················· | ····················· | ···················· | ···················· | ···················· |
| K | 1476 | hCoV-19/Australia/VIC7519/2020\|EPI_ISL_564465\|2020 | ···················· | ···················· | ····················· | ···················· | ···················· | ···················· |
| K | 1477 | hCoV-19/Australia/VIC7520/2020\|EPI_ISL_564466\|2020 | ···················· | ···················· | ····················· | ···················· | ···················· | ···················· |
| K | 1479 | hCoV-19/Australia/VIC9813/2020\|EPI_ISL_565732\|2020 | ···················· | ···················· | ···K················· | ···················· | ···················· | ···················· |
| K | 1480 | hCoV-19/Australia/VIC9816/2020\|EPI_ISL_565733\|2020 | ···················· | ···················· | ····················· | ···················· | ···················· | ···················· |
| K | 1481 | hCoV-19/Australia/VIC9824/2020\|EPI_ISL_565734\|2020 | ···················· | ···················· | ····················· | ···················· | ···················· | ···················· |
| K | 1482 | hCoV-19/Australia/VIC9827/2020\|EPI_ISL_565735\|2020 | ···················· | ···················· | ···K················· | ···················· | ···················· | ···················· |
| K | 1483 | hCoV-19/Australia/VIC9832/2020\|EPI_ISL_565736\|2020 | ···················· | ···················· | ····················· | ···················· | ···················· | ···················· |
| K | 1484 | hCoV-19/Australia/VIC9833/2020\|EPI_ISL_565737\|2020 | ···················· | ···················· | ····················· | ···················· | ···················· | ···················· |
| K | 1485 | hCoV-19/Australia/VIC9835/2020\|EPI_ISL_565738\|2020 | ···················· | ···················· | ····················· | ···················· | ···················· | ···················· |
| K | 1486 | hCoV-19/Australia/VIC9837/2020\|EPI_ISL_565739\|2020 | ···················· | ···················· | ····················· | ···················· | ···················· | ···················· |
| K | 1487 | hCoV-19/Australia/VIC9838/2020\|EPI_ISL_565740\|2020 | ···················· | ···················· | ····················· | ···················· | ···················· | ···················· |
| K | 1488 | hCoV-19/Australia/VIC9839/2020\|EPI_ISL_565741\|2020 | ···················· | ···················· | ····················· | ···················· | ···················· | ···················· |
| K | 1489 | hCoV-19/Australia/VIC9851/2020\|EPI_ISL_565742\|2020 | ···················· | ···················· | ····················· | ···················· | ···················· | ···················· |
| K | 1490 | hCoV-19/Australia/VIC9857/2020\|EPI_ISL_565743\|2020 | ···················· | ···················· | ····················· | ···················· | ···················· | ···················· |
| K | 1491 | hCoV-19/Australia/VIC9858/2020\|EPI_ISL_565744\|2020 | ···················· | ···················· | ····················· | ···················· | ···················· | ···················· |
| K | 1492 | hCoV-19/Australia/VIC9860/2020\|EPI_ISL_565745\|2020 | ···················· | ······A············· | ····················· | ···················· | ···················· | ···················· |
| K | 1493 | hCoV-19/Australia/VIC9861/2020\|EPI_ISL_565746\|2020 | ···················· | ···················· | ····················· | ···················· | ···················· | ···················· |
| K | 1494 | hCoV-19/Australia/VIC9862/2020\|EPI_ISL_565747\|2020 | ···················· | ···················· | ····················· | ···················· | ···················· | ···················· |
| K | 1495 | hCoV-19/Australia/VIC9864/2020\|EPI_ISL_565748\|2020 | ···················· | ···················· | ····················· | ···················· | ···················· | ···················· |
| K | 1496 | hCoV-19/Australia/VIC9865/2020\|EPI_ISL_565749\|2020 | ···················· | ···················· | ····················· | ···················· | ···················· | ···················· |
| K | 1497 | hCoV-19/Australia/VIC9866/2020\|EPI_ISL_565750\|2020 | ···················· | ···················· | ····················· | ···················· | ···················· | ···················· |
| K | 1499 | hCoV-19/Australia/VIC9870/2020\|EPI_ISL_565752\|2020 | ···················· | ···················· | ····················· | ···················· | ···················· | ···················· |
| K | 1500 | hCoV-19/Australia/VIC9871/2020\|EPI_ISL_565753\|2020 | ···················· | ···················· | ····················· | ···················· | ···················· | ···················· |
| K | 1501 | hCoV-19/Australia/VIC9872/2020\|EPI_ISL_565754\|2020 | ···················· | ···················· | ····················· | ···················· | ···················· | ···················· |
| K | 1502 | hCoV-19/Australia/VIC9873/2020\|EPI_ISL_565755\|2020 | ···················· | ···················· | ····················· | ···················· | ···················· | ···················· |
| K | 1503 | hCoV-19/Australia/VIC9874/2020\|EPI_ISL_565756\|2020 | ···················· | ···················· | ····················· | ···················· | ···················· | ···················· |
| K | 1504 | hCoV-19/Australia/VIC9875/2020\|EPI_ISL_565757\|2020 | ···················· | ···················· | ····················· | ···················· | ···················· | ···················· |
| K | 1505 | hCoV-19/Australia/VIC9877/2020\|EPI_ISL_565758\|2020 | ···················· | ···················· | ····················· | ···················· | ···················· | ···················· |
| K | 1506 | hCoV-19/Australia/VIC9878/2020\|EPI_ISL_565759\|2020 | ···················· | ···················· | ····················· | ···················· | ···················· | ···················· |
| K | 1507 | hCoV-19/Australia/VIC9879/2020\|EPI_ISL_565760\|2020 | ···················· | ···················· | ····················· | ···················· | ···················· | ···················· |
| K | 1508 | hCoV-19/Australia/VIC9880/2020\|EPI_ISL_565761\|2020 | ···················· | ···················· | ····················· | ···················· | ···················· | ···················· |
| K | 1509 | hCoV-19/Australia/VIC9881/2020\|EPI_ISL_565762\|2020 | ···················· | ···················· | ····················· | ···················· | ···················· | ···················· |
| K | 1510 | hCoV-19/Australia/VIC9883/2020\|EPI_ISL_565763\|2020 | ···················· | ···················· | ····················· | ···················· | ···················· | ···················· |
| K | 1511 | hCoV-19/Australia/VIC9889/2020\|EPI_ISL_565764\|2020 | ···················· | ···················· | ····················· | ···················· | ···················· | ···················· |

| | | | | | | | |
|---|---|---|---|---|---|---|---|
| K | 1512 hCoV-19/Australia/VIC9890/2020\|EPI_ISL_565765\|2020 | .................... | .................... | .................... | .................... | .................... | .................... |
| K | 1513 hCoV-19/Australia/VIC9892/2020\|EPI_ISL_565766\|2020 | .................... | .................... | .................... | .................... | .................... | .................... |
| K | 1514 hCoV-19/Australia/VIC9893/2020\|EPI_ISL_565767\|2020 | .................... | .................... | .................... | .................... | .................... | .................... |
| K | 1515 hCoV-19/Australia/VIC9895/2020\|EPI_ISL_565768\|2020 | .................... | .................... | .................... | .................... | .................... | .................... |
| K | 1516 hCoV-19/Australia/VIC9898/2020\|EPI_ISL_565769\|2020 | .................... | .................... | .................... | .................... | .................... | .................... |
| K | 1517 hCoV-19/Australia/VIC9899/2020\|EPI_ISL_565770\|2020 | .................... | .................... | .................... | .................... | .................... | .................... |
| K | 1518 hCoV-19/Australia/VIC9900/2020\|EPI_ISL_565771\|2020 | .................... | .................... | .................... | .................... | .................... | .................... |
| K | 1519 hCoV-19/Australia/VIC9902/2020\|EPI_ISL_565772\|2020 | .................... | .................... | .................... | .................... | .................... | .................... |
| K | 1520 hCoV-19/Australia/VIC9903/2020\|EPI_ISL_565773\|2020 | .................... | .................... | .................... | .................... | .................... | .................... |
| K | 1521 hCoV-19/Australia/VIC9904/2020\|EPI_ISL_565774\|2020 | .................... | .................... | .................... | .................... | .................... | .................... |
| K | 1522 hCoV-19/Australia/VIC9906/2020\|EPI_ISL_565775\|2020 | .................... | .................... | .................... | .................... | .................... | .................... |
| K | 1523 hCoV-19/Australia/VIC9907/2020\|EPI_ISL_565776\|2020 | .................... | .................... | .................... | .................... | .................... | .................... |
| K | 1524 hCoV-19/Australia/VIC9908/2020\|EPI_ISL_565777\|2020 | .................... | .................... | .................... | .................... | .................... | .................... |
| K | 1525 hCoV-19/Australia/VIC9917/2020\|EPI_ISL_565779\|2020 | .................... | .................... | .................... | .................... | .................... | .................... |
| K | 1526 hCoV-19/Australia/VIC9936/2020\|EPI_ISL_565787\|2020 | .................... | .................... | .................... | .................... | .................... | .................... |
| K | 1527 hCoV-19/Australia/VIC9937/2020\|EPI_ISL_565788\|2020 | .................... | .................... | .................... | .................... | .................... | .................... |
| K | 1528 hCoV-19/Australia/VIC9938/2020\|EPI_ISL_565789\|2020 | .................... | .................... | .................... | .................... | .................... | .................... |
| K | 1529 hCoV-19/Australia/VIC9939/2020\|EPI_ISL_565790\|2020 | .................... | .................... | .................... | .................... | .................... | .................... |
| K | 1530 hCoV-19/Australia/VIC9941/2020\|EPI_ISL_565791\|2020 | .................... | .................... | .................... | .................... | .................... | .................... |
| K | 1531 hCoV-19/Australia/VIC9943/2020\|EPI_ISL_565792\|2020 | .................... | .................... | .................... | .................... | .................... | .................... |
| K | 1532 hCoV-19/Australia/VIC9944/2020\|EPI_ISL_565793\|2020 | .................... | .................... | .................... | .................... | .................... | .................... |
| K | 1533 hCoV-19/Australia/VIC9945/2020\|EPI_ISL_565794\|2020 | .................... | .................... | .................... | .................... | .................... | .................... |
| K | 1534 hCoV-19/Australia/VIC9946/2020\|EPI_ISL_565795\|2020 | .................... | .................... | .................... | .................... | .................... | .................... |
| K | 1535 hCoV-19/Australia/VIC9947/2020\|EPI_ISL_565796\|2020 | .................... | .................... | .................... | .................... | .................... | .................... |
| K | 1536 hCoV-19/Australia/VIC9948/2020\|EPI_ISL_565797\|2020 | .................... | .................... | .................... | .................... | .................... | .................... |
| K | 1537 hCoV-19/England/GSTT-6731/2020\|EPI_ISL_560722\|2020 | .................... | .................... | .................... | .................... | .................... | .................... |
| K | 1538 hCoV-19/England/GSTT-6732/2020\|EPI_ISL_560723\|2020 | .................... | .................... | .................... | .................... | .................... | .................... |
| K | 1539 hCoV-19/England/GSTT-6742/2020\|EPI_ISL_560724\|2020 | .................... | .................... | .................... | .................... | .................... | .................... |
| K | 1540 hCoV-19/England/GSTT-6744/2020\|EPI_ISL_560725\|2020 | .................... | .................... | .................... | .................... | .................... | .................... |
| K | 1541 hCoV-19/England/GSTT-6746/2020\|EPI_ISL_560726\|2020 | .................... | .................... | .................... | .................... | .................... | .................... |
| K | 1542 hCoV-19/England/GSTT-6749/2020\|EPI_ISL_560727\|2020 | .................... | .................... | .................... | .................... | .................... | .................... |
| K | 1543 hCoV-19/England/GSTT-6754/2020\|EPI_ISL_560729\|2020 | .................... | .................... | .................... | .................... | .................... | .................... |
| K | 1544 hCoV-19/England/GSTT-6757/2020\|EPI_ISL_560730\|2020 | .................... | .................... | .................... | .................... | .................... | .................... |
| K | 1545 hCoV-19/England/GSTT-6767/2020\|EPI_ISL_560731\|2020 | .................... | .................... | .................... | .................... | .................... | .................... |
| K | 1546 hCoV-19/England/GSTT-6769/2020\|EPI_ISL_560733\|2020 | .................... | .................... | .................... | .................... | .................... | .................... |
| K | 1547 hCoV-19/England/GSTT-6773/2020\|EPI_ISL_560734\|2020 | .................... | .................... | .................... | .................... | .................... | .................... |
| K | 1548 hCoV-19/England/GSTT-6776/2020\|EPI_ISL_560736\|2020 | .................... | .................... | .................... | .................... | .................... | .................... |
| K | 1549 hCoV-19/England/GSTT-6784/2020\|EPI_ISL_560737\|2020 | .................... | .................... | .................... | .................... | .................... | .................... |
| K | 1550 hCoV-19/England/GSTT-6843/2020\|EPI_ISL_560739\|2020 | .................... | .................... | .................... | .................... | .................... | .................... |
| K | 1551 hCoV-19/England/GSTT-6885/2020\|EPI_ISL_560740\|2020 | .................... | .................... | .................... | .................... | .................... | .................... |
| K | 1552 hCoV-19/USA/DE-DHSS-F917628/2020\|EPI_ISL_560741\|20 | .................... | .................... | .................... | .................... | .................... | .................... |
| K | 1553 hCoV-19/USA/MN-MDH-1656/2020\|EPI_ISL_560743\|2020-0 | .................... | .................... | .................... | .................... | .................... | .................... |
| K | 1554 hCoV-19/USA/MN-MDH-1657/2020\|EPI_ISL_560744\|2020-0 | .................... | .................... | .................... | .................... | .................... | .................... |
| K | 1555 hCoV-19/USA/MN-MDH-1658/2020\|EPI_ISL_560745\|2020-0 | .................... | .................... | .................... | .................... | .................... | .................... |
| K | 1556 hCoV-19/USA/MN-MDH-1659/2020\|EPI_ISL_560746\|2020-0 | .................... | ....G............... | .................... | .................... | .................... | .................... |
| K | 1557 hCoV-19/USA/MN-MDH-1660/2020\|EPI_ISL_560747\|2020-0 | .................... | ....G............... | .................... | .................... | .................... | .................... |
| K | 1558 hCoV-19/USA/MN-MDH-1661/2020\|EPI_ISL_560748\|2020-0 | .................... | .................... | .................... | .................... | .................... | .................... |
| K | 1559 hCoV-19/USA/MN-MDH-1662/2020\|EPI_ISL_560749\|2020-0 | .................... | .................... | .................... | .................... | .................... | .................... |
| K | 1560 hCoV-19/USA/MN-MDH-1663/2020\|EPI_ISL_560750\|2020-0 | .................... | .................... | .................... | .................... | .................... | .................... |
| K | 1561 hCoV-19/USA/MN-MDH-1664/2020\|EPI_ISL_560751\|2020-0 | .................... | .................... | .................... | .................... | .................... | .................... |
| K | 1562 hCoV-19/USA/MN-MDH-1665/2020\|EPI_ISL_560752\|2020-0 | .................... | .................... | .................... | .................... | .................... | .................... |
| K | 1563 hCoV-19/USA/MN-MDH-1666/2020\|EPI_ISL_560753\|2020-0 | .................... | .................... | .................... | .................... | .................... | .................... |
| K | 1564 hCoV-19/USA/MN-MDH-1667/2020\|EPI_ISL_560754\|2020-0 | .................... | .................... | .................... | .................... | .................... | .................... |
| K | 1565 hCoV-19/USA/MN-MDH-1668/2020\|EPI_ISL_560755\|2020-0 | .................... | .................... | .................... | .................... | .................... | .................... |
| K | 1566 hCoV-19/USA/MN-MDH-1669/2020\|EPI_ISL_560756\|2020-0 | .................... | .................... | .................... | .................... | .................... | .................... |
| K | 1567 hCoV-19/USA/MN-MDH-1670/2020\|EPI_ISL_560757\|2020-0 | .................... | .................... | .................... | .................... | .................... | .................... |
| K | 1568 hCoV-19/USA/MN-MDH-1671/2020\|EPI_ISL_560758\|2020-0 | .................... | .................... | .................... | .................... | .................... | .................... |
| K | 1569 hCoV-19/USA/MN-MDH-1672/2020\|EPI_ISL_560759\|2020-0 | .................... | .................... | .................... | .................... | .................... | .................... |
| K | 1570 hCoV-19/USA/MN-MDH-1673/2020\|EPI_ISL_560760\|2020-0 | .................... | .................... | .................... | .................... | .................... | .................... |
| K | 1571 hCoV-19/USA/MN-MDH-1674/2020\|EPI_ISL_560761\|2020-0 | .................... | ....G............... | .................... | .................... | .................... | .................... |
| K | 1572 hCoV-19/USA/MN-MDH-1675/2020\|EPI_ISL_560762\|2020-0 | .................... | .................... | .................... | .................... | .................... | .................... |
| K | 1573 hCoV-19/USA/MN-MDH-1676/2020\|EPI_ISL_560763\|2020-0 | .................... | .................... | .................... | .................... | .................... | .................... |
| K | 1574 hCoV-19/USA/MN-MDH-1677/2020\|EPI_ISL_560764\|2020-0 | .................... | .................... | .................... | .................... | .................... | .................... |
| K | 1575 hCoV-19/USA/MN-MDH-1678/2020\|EPI_ISL_560765\|2020-0 | .................... | .................... | .................... | .................... | .................... | .................... |
| K | 1576 hCoV-19/USA/MN-MDH-1679/2020\|EPI_ISL_560766\|2020-0 | .................... | .................... | .................... | .................... | .................... | .................... |
| K | 1577 hCoV-19/USA/MN-MDH-1680/2020\|EPI_ISL_560767\|2020-0 | .................... | .................... | .................... | .................... | .................... | .................... |
| K | 1578 hCoV-19/USA/MN-MDH-1681/2020\|EPI_ISL_560768\|2020-0 | .................... | .................... | .................... | .................... | .................... | .................... |
| K | 1579 hCoV-19/USA/MN-MDH-1682/2020\|EPI_ISL_560769\|2020-0 | .................... | .................... | .................... | .................... | .................... | .................... |
| K | 1580 hCoV-19/USA/MN-MDH-1684/2020\|EPI_ISL_560771\|2020-0 | .................... | .................... | .................... | .................... | .................... | .................... |
| K | 1581 hCoV-19/USA/MN-MDH-1685/2020\|EPI_ISL_560772\|2020-0 | .................... | .................... | .................... | .................... | .................... | .................... |
| K | 1582 hCoV-19/USA/MN-MDH-1686/2020\|EPI_ISL_560773\|2020-0 | .................... | .................... | .................... | .................... | .................... | .................... |
| K | 1583 hCoV-19/USA/MN-MDH-1687/2020\|EPI_ISL_560774\|2020-0 | .................... | .................... | .................... | .................... | .................... | .................... |
| K | 1584 hCoV-19/USA/MN-MDH-1688/2020\|EPI_ISL_560775\|2020-0 | .................... | .................... | .................... | .................... | .................... | .................... |
| K | 1585 hCoV-19/USA/MN-MDH-1689/2020\|EPI_ISL_560776\|2020-0 | .................... | .................... | .................... | .................... | .................... | .................... |
| K | 1586 hCoV-19/USA/MN-MDH-1690/2020\|EPI_ISL_560777\|2020-0 | .................... | .................... | .................... | .................... | .................... | .................... |
| K | 1587 hCoV-19/USA/MN-MDH-1691/2020\|EPI_ISL_560778\|2020-0 | .................... | .................... | .................... | .................... | .................... | .................... |
| K | 1588 hCoV-19/USA/MN-MDH-1692/2020\|EPI_ISL_560779\|2020-0 | .................... | .................... | .................... | .................... | .................... | .................... |
| K | 1589 hCoV-19/USA/MN-MDH-1693/2020\|EPI_ISL_560780\|2020-0 | .................... | .................... | .................... | .................... | .................... | .................... |
| K | 1590 hCoV-19/USA/MN-MDH-1694/2020\|EPI_ISL_560781\|2020-0 | .................... | .................... | .................... | .................... | .................... | .................... |
| K | 1591 hCoV-19/Australia/VIC10831/2020\|EPI_ISL_563116\|202 | .................... | .................... | .................... | .................... | .................... | .................... |

| | | | | | | | | |
|---|---|---|---|---|---|---|---|---|
| K | 1592 hCoV-19/Australia/VIC10832/2020\|EPI_ISL_563117\|202 | ···················· | ···················· | ···················· | ···················· | ···················· | ···················· | |
| K | 1593 hCoV-19/Australia/VIC10836/2020\|EPI_ISL_563120\|202 | ···················· | ···················· | ···················· | ···················· | ···················· | ···················· | |
| K | 1594 hCoV-19/Australia/VIC10838/2020\|EPI_ISL_563121\|202 | ···················· | ···················· | ···················· | ···················· | ···················· | ···················· | |
| K | 1595 hCoV-19/Australia/VIC10840/2020\|EPI_ISL_563122\|202 | ···················· | ···················· | ···················· | ···················· | ···················· | ···················· | |
| K | 1596 hCoV-19/Australia/VIC10841/2020\|EPI_ISL_563123\|202 | ···················· | ···················· | ···················· | ···················· | ···················· | ···················· | |
| K | 1597 hCoV-19/Australia/VIC10847/2020\|EPI_ISL_563124\|202 | ···················· | ···················· | ···················· | ···················· | ···················· | ···················· | |
| K | 1598 hCoV-19/Australia/VIC10882/2020\|EPI_ISL_563136\|202 | ···················· | ···················· | ··K················· | ···················· | ···················· | ···················· | |
| K | 1599 hCoV-19/Australia/VIC10883/2020\|EPI_ISL_563137\|202 | ···················· | ···················· | ···················· | ···················· | ···················· | ···················· | |
| K | 1601 hCoV-19/Australia/VIC10888/2020\|EPI_ISL_563139\|202 | ···················· | ···················· | ···················· | ···················· | ···················· | ···················· | |
| K | 1602 hCoV-19/Australia/VIC10889/2020\|EPI_ISL_563140\|202 | ···················· | ···················· | ···················· | ···················· | ···················· | ···················· | |
| K | 1603 hCoV-19/Australia/VIC10890/2020\|EPI_ISL_563141\|202 | ···················· | ···················· | ···················· | ···················· | ···················· | ···················· | |
| K | 1604 hCoV-19/Australia/VIC10891/2020\|EPI_ISL_563142\|202 | ···················· | ···················· | ···················· | ···················· | ···················· | ···················· | |
| K | 1605 hCoV-19/Australia/VIC10896/2020\|EPI_ISL_563143\|202 | ···················· | ···················· | ··K················· | ···················· | ···················· | ···················· | |
| K | 1606 hCoV-19/Australia/VIC10897/2020\|EPI_ISL_563144\|202 | ···················· | ···················· | ···················· | ···················· | ···················· | ···················· | |
| K | 1608 hCoV-19/Australia/VIC10903/2020\|EPI_ISL_563146\|202 | ···················· | ···················· | ···················· | ···················· | ···················· | ···················· | |
| K | 1610 hCoV-19/Australia/VIC10905/2020\|EPI_ISL_563148\|202 | ···················· | ···················· | ···················· | ···················· | ···················· | ···················· | |
| K | 1612 hCoV-19/Australia/VIC10910/2020\|EPI_ISL_563150\|202 | ···················· | ···················· | ···················· | ···················· | ···················· | ···················· | |
| K | 1615 hCoV-19/Australia/VIC10917/2020\|EPI_ISL_563153\|202 | ···················· | ···················· | ···················· | ···················· | ···················· | ···················· | |
| K | 1616 hCoV-19/Australia/VIC10923/2020\|EPI_ISL_563154\|202 | ···················· | ···················· | ···················· | ···················· | ···················· | ···················· | |
| K | 1617 hCoV-19/Australia/VIC10925/2020\|EPI_ISL_563155\|202 | ···················· | ···················· | ···················· | ···················· | ···················· | ···················· | |
| K | 1618 hCoV-19/Australia/VIC10926/2020\|EPI_ISL_563156\|202 | ···················· | ···················· | ···················· | ···················· | ···················· | ···················· | |
| K | 1619 hCoV-19/Australia/VIC10928/2020\|EPI_ISL_563157\|202 | ···················· | ···················· | ···················· | ···················· | ···················· | ···················· | |
| K | 1620 hCoV-19/Australia/VIC7424/2020\|EPI_ISL_564400\|2020 | ···················· | ···················· | ···················· | ···················· | ···················· | ···················· | |
| K | 1621 hCoV-19/Australia/VIC7495/2020\|EPI_ISL_564448\|2020 | ···················· | ···················· | ···················· | ···················· | ···················· | ···················· | |
| K | 1622 hCoV-19/Australia/VIC7497/2020\|EPI_ISL_564450\|2020 | ···················· | ···················· | ···················· | ···················· | ···················· | ···················· | |
| K | 1623 hCoV-19/Australia/VIC7499/2020\|EPI_ISL_564451\|2020 | ···················· | ···················· | ···················· | ···················· | ···················· | ···················· | |
| K | 1624 hCoV-19/Australia/VIC7502/2020\|EPI_ISL_564452\|2020 | ···················· | ···················· | ···················· | ···················· | ···················· | ···················· | |
| K | 1625 hCoV-19/Australia/VIC7504/2020\|EPI_ISL_564453\|2020 | ···················· | ···················· | ···················· | ···················· | ···················· | ···················· | |
| K | 1626 hCoV-19/Australia/VIC7505/2020\|EPI_ISL_564454\|2020 | ···················· | ···················· | ···················· | ···················· | ···················· | ···················· | |
| K | 1627 hCoV-19/Australia/VIC7506/2020\|EPI_ISL_564455\|2020 | ···················· | ······A············· | ···················· | ···················· | ···················· | ···················· | |
| K | 1628 hCoV-19/Australia/VIC7521/2020\|EPI_ISL_564467\|2020 | ···················· | ···················· | ···················· | ···················· | ···················· | ···················· | |
| K | 1629 hCoV-19/Australia/VIC7522/2020\|EPI_ISL_564468\|2020 | ···················· | ···················· | ···················· | ···················· | ···················· | ···················· | |
| K | 1630 hCoV-19/Australia/VIC7523/2020\|EPI_ISL_564469\|2020 | ···················· | ···················· | ···················· | ···················· | ···················· | ···················· | |
| K | 1631 hCoV-19/Australia/VIC7525/2020\|EPI_ISL_564470\|2020 | ···················· | ···················· | ···················· | ···················· | ···················· | ···················· | |
| K | 1632 hCoV-19/Australia/VIC7527/2020\|EPI_ISL_564471\|2020 | ···················· | ···················· | ···················· | ···················· | ···················· | ···················· | |
| K | 1633 hCoV-19/Australia/VIC7528/2020\|EPI_ISL_564472\|2020 | ···················· | ···················· | ···················· | ···················· | ···················· | ···················· | |
| K | 1634 hCoV-19/Australia/VIC7529/2020\|EPI_ISL_564473\|2020 | ···················· | ···················· | ···················· | ···················· | ···················· | ···················· | |
| K | 1635 hCoV-19/Australia/VIC7530/2020\|EPI_ISL_564474\|2020 | ···················· | ···················· | ···················· | ···················· | ···················· | ···················· | |
| K | 1636 hCoV-19/Australia/VIC7531/2020\|EPI_ISL_564475\|2020 | ···················· | ···················· | ···················· | ···················· | ···················· | ···················· | |
| K | 1637 hCoV-19/Australia/VIC7532/2020\|EPI_ISL_564476\|2020 | ···················· | ···················· | ···················· | ···················· | ···················· | ···················· | |
| K | 1638 hCoV-19/Australia/VIC7533/2020\|EPI_ISL_564477\|2020 | ···················· | ···················· | ···················· | ···················· | ···················· | ···················· | |
| K | 1639 hCoV-19/Australia/VIC7534/2020\|EPI_ISL_564478\|2020 | ···················· | ···················· | ···················· | ···················· | ···················· | ···················· | |
| K | 1640 hCoV-19/Australia/VIC7535/2020\|EPI_ISL_564479\|2020 | ···················· | ···················· | ···················· | ···················· | ···················· | ···················· | |
| K | 1641 hCoV-19/Australia/VIC7536/2020\|EPI_ISL_564480\|2020 | ···················· | ···················· | ···················· | ···················· | ···················· | ···················· | |
| K | 1642 hCoV-19/Australia/VIC7537/2020\|EPI_ISL_564481\|2020 | ···················· | ···················· | ···················· | ···················· | ···················· | ···················· | |
| K | 1643 hCoV-19/Australia/VIC7538/2020\|EPI_ISL_564482\|2020 | ···················· | ···················· | ···················· | ···················· | ···················· | ···················· | |
| K | 1644 hCoV-19/Australia/VIC7539/2020\|EPI_ISL_564483\|2020 | ···················· | ···················· | ···················· | ···················· | ···················· | ···················· | |
| K | 1645 hCoV-19/Australia/VIC7540/2020\|EPI_ISL_564484\|2020 | ···················· | ···················· | ···················· | ···················· | ···················· | ···················· | |
| K | 1646 hCoV-19/Australia/VIC7542/2020\|EPI_ISL_564485\|2020 | ···················· | ···················· | ···················· | ···················· | ···················· | ···················· | |
| K | 1647 hCoV-19/Australia/VIC7543/2020\|EPI_ISL_564486\|2020 | ···················· | ···················· | ···················· | ···················· | ···················· | ···················· | |
| K | 1648 hCoV-19/Australia/VIC7544/2020\|EPI_ISL_564487\|2020 | ···················· | ···················· | ···················· | ···················· | ···················· | ···················· | |
| K | 1649 hCoV-19/Australia/VIC7545/2020\|EPI_ISL_564488\|2020 | ···················· | ···················· | ···················· | ···················· | ···················· | ···················· | |
| K | 1650 hCoV-19/Australia/VIC9764/2020\|EPI_ISL_565700\|2020 | ···················· | ···················· | ···················· | ···················· | NNN················· | ···················· | 1 |
| K | 1652 hCoV-19/Australia/VIC9766/2020\|EPI_ISL_565702\|2020 | ···················· | ···················· | ···················· | ···················· | ···················· | ···················· | |
| K | 1655 hCoV-19/Australia/VIC9772/2020\|EPI_ISL_565705\|2020 | ···················· | ···················· | ···················· | ···················· | ···················· | ···················· | |
| K | 1657 hCoV-19/Australia/VIC9775/2020\|EPI_ISL_565707\|2020 | ···················· | ···················· | ···················· | ···················· | ···················· | ···················· | |
| K | 1658 hCoV-19/Australia/VIC9777/2020\|EPI_ISL_565708\|2020 | ···················· | ···················· | ···················· | ···················· | ···················· | ···················· | |
| K | 1659 hCoV-19/Australia/VIC9778/2020\|EPI_ISL_565709\|2020 | ···················· | ···················· | ···················· | ···················· | ···················· | ···················· | |
| K | 1660 hCoV-19/Australia/VIC9780/2020\|EPI_ISL_565710\|2020 | ···················· | ···················· | ···················· | ···················· | ···················· | ···················· | |
| K | 1661 hCoV-19/Australia/VIC9781/2020\|EPI_ISL_565711\|2020 | ···················· | ···················· | ···················· | ···················· | ···················· | ···················· | |
| K | 1662 hCoV-19/Australia/VIC9783/2020\|EPI_ISL_565712\|2020 | ···················· | ···················· | ···················· | ···················· | ···················· | ···················· | |
| K | 1663 hCoV-19/Australia/VIC9784/2020\|EPI_ISL_565713\|2020 | ···················· | ···················· | ···················· | ···················· | ···················· | ···················· | |
| K | 1664 hCoV-19/Australia/VIC9785/2020\|EPI_ISL_565714\|2020 | ···················· | ···················· | ···················· | ···················· | ···················· | ···················· | |
| K | 1666 hCoV-19/Australia/VIC9787/2020\|EPI_ISL_565716\|2020 | ···················· | ···················· | ···················· | ···················· | ···················· | ···················· | |
| K | 1667 hCoV-19/Australia/VIC9788/2020\|EPI_ISL_565717\|2020 | ···················· | ···················· | ···················· | ···················· | ···················· | ···················· | |
| K | 1669 hCoV-19/Australia/VIC9791/2020\|EPI_ISL_565719\|2020 | ···················· | ···················· | ···················· | ···················· | ···················· | ···················· | |
| K | 1670 hCoV-19/Australia/VIC9792/2020\|EPI_ISL_565720\|2020 | ···················· | ···················· | ···················· | ···················· | ···················· | ···················· | |
| K | 1671 hCoV-19/Australia/VIC9793/2020\|EPI_ISL_565721\|2020 | ···················· | ···················· | ···················· | ···················· | ···················· | ···················· | |
| K | 1672 hCoV-19/Australia/VIC9796/2020\|EPI_ISL_565722\|2020 | ···················· | ···················· | ···················· | ···················· | ···················· | ···················· | |
| K | 1673 hCoV-19/Australia/VIC9798/2020\|EPI_ISL_565723\|2020 | ···················· | ···················· | ···················· | ···················· | ···················· | ···················· | |
| K | 1674 hCoV-19/Australia/VIC9800/2020\|EPI_ISL_565724\|2020 | ···················· | ···················· | ···················· | ···················· | ···················· | ···················· | |
| K | 1677 hCoV-19/Australia/VIC9805/2020\|EPI_ISL_565727\|2020 | ···················· | ···················· | ···················· | ···················· | ···················· | ···················· | |
| K | 1679 hCoV-19/Australia/VIC9808/2020\|EPI_ISL_565729\|2020 | ···················· | ···················· | ···················· | ···················· | ···················· | ···················· | |
| K | 1680 hCoV-19/Australia/VIC9811/2020\|EPI_ISL_565730\|2020 | ···················· | ···················· | ··K················· | ···················· | ···················· | ···················· | |
| K | 1681 hCoV-19/Australia/VIC9812/2020\|EPI_ISL_565731\|2020 | ···················· | ···················· | ··K················· | ···················· | ···················· | ···················· | |
| K | 1682 hCoV-19/Australia/VIC9909/2020\|EPI_ISL_565778\|2020 | ···················· | ···················· | ···················· | ···················· | ···················· | ···················· | |
| K | 1683 hCoV-19/Australia/VIC9918/2020\|EPI_ISL_565780\|2020 | ···················· | ···················· | ···················· | ···················· | ···················· | ···················· | |
| K | 1684 hCoV-19/Australia/VIC9921/2020\|EPI_ISL_565781\|2020 | ···················· | ···················· | ···················· | ···················· | ···················· | ···················· | |
| K | 1685 hCoV-19/Australia/VIC9922/2020\|EPI_ISL_565782\|2020 | ···················· | ···················· | ···················· | ···················· | ···················· | ···················· | |
| K | 1686 hCoV-19/Australia/VIC9925/2020\|EPI_ISL_565783\|2020 | ···················· | ···················· | ···················· | ···················· | ···················· | ···················· | |

| | | | | | | | | |
|---|---|---|---|---|---|---|---|---|
| K | 1687 | hCoV-19/Australia/VIC9927/2020\|EPI_ISL_565784\|2020 | ···················· | ···················· | ····················· | ···················· | ···················· | ···················· |
| K | 1688 | hCoV-19/Australia/VIC9934/2020\|EPI_ISL_565785\|2020 | ···················· | ···················· | ····················· | ···················· | ···················· | ···················· |
| K | 1689 | hCoV-19/Australia/VIC9935/2020\|EPI_ISL_565786\|2020 | ···················· | ···················· | ····················· | ···················· | ···················· | ···················· |
| K | 1690 | hCoV-19/Australia/VIC9949/2020\|EPI_ISL_565798\|2020 | ···················· | ···················· | ····················· | ···················· | ···················· | ···················· |
| K | 1691 | hCoV-19/Australia/VIC9951/2020\|EPI_ISL_565799\|2020 | ···················· | ···················· | ····················· | ···················· | ···················· | ···················· |
| K | 1692 | hCoV-19/France/PAC-IHU-1277/2020\|EPI_ISL_569105\|20 | ···················· | ·A·················· | ····················· | ···················· | ···················· | ···················· |
| K | 1693 | hCoV-19/France/PAC-IHU-1283/2020\|EPI_ISL_569107\|20 | ···················· | ·A·················· | ····················· | ···················· | ···················· | ···················· |
| K | 1694 | hCoV-19/France/PAC-IHU-1289/2020\|EPI_ISL_569108\|20 | ···················· | ···················· | ····················· | ···················· | ···················· | ···················· |
| K | 1695 | hCoV-19/France/PAC-IHU-1292/2020\|EPI_ISL_569109\|20 | ···················· | ···················· | ····················· | ···················· | ···················· | ···················· |
| K | 1696 | hCoV-19/France/PAC-IHU-1295/2020\|EPI_ISL_569110\|20 | ···················· | ···················· | ····················· | ···················· | ···················· | ···················· |
| K | 1697 | hCoV-19/France/PAC-IHU-1296/2020\|EPI_ISL_569111\|20 | ···················· | ···················· | ····················· | ···················· | ···················· | ···················· |
| K | 1698 | hCoV-19/France/PAC-IHU-1307/2020\|EPI_ISL_569113\|20 | ···················· | ···················· | ····················· | ···················· | ···················· | ···················· |
| K | 1699 | hCoV-19/France/PAC-IHU-1315/2020\|EPI_ISL_569114\|20 | ···················· | ·A·················· | ····················· | ···················· | ···················· | ···················· |
| K | 1700 | hCoV-19/France/PAC-IHU-1316/2020\|EPI_ISL_569115\|20 | ···················· | ···················· | ····················· | ···················· | ···················· | ···················· |
| K | 1701 | hCoV-19/France/PAC-IHU-1325/2020\|EPI_ISL_569116\|20 | ···················· | ···················· | ····················· | ···················· | ···················· | ···················· |
| K | 1702 | hCoV-19/France/PAC-IHU-1327/2020\|EPI_ISL_569117\|20 | ········T··········· | ···················· | ····················· | ···················· | ···················· | ···················· |
| K | 1703 | hCoV-19/France/PAC-IHU-1329/2020\|EPI_ISL_569119\|20 | ···················· | ···················· | ····················· | ···················· | ···················· | ···················· |
| K | 1704 | hCoV-19/France/PAC-IHU-0037/2020\|EPI_ISL_569261\|20 | ···················· | ···················· | ····················· | ···················· | ···················· | ···················· |
| K | 1705 | hCoV-19/France/PAC-IHU-0038/2020\|EPI_ISL_569262\|20 | ···················· | ···················· | ····················· | ···················· | ···················· | ···················· |
| K | 1706 | hCoV-19/France/PAC-IHU-0039/2020\|EPI_ISL_569263\|20 | ···················· | ···················· | ····················· | ···················· | ···················· | ···················· |
| K | 1707 | hCoV-19/France/PAC-IHU-0040/2020\|EPI_ISL_569264\|20 | ···················· | ···················· | ····················· | ···················· | ···················· | ···················· |
| K | 1708 | hCoV-19/France/PAC-IHU-0044/2020\|EPI_ISL_569265\|20 | ···················· | ···················· | ····················· | ···················· | ···················· | ···················· |
| K | 1709 | hCoV-19/France/PAC-IHU-0045/2020\|EPI_ISL_569266\|20 | ···················· | ···················· | ····················· | ···················· | ···················· | ···················· |
| K | 1710 | hCoV-19/France/PAC-IHU-0046/2020\|EPI_ISL_569267\|20 | ···················· | ···················· | ····················· | ···················· | ···················· | ···················· |
| K | 1711 | hCoV-19/France/PAC-IHU-0047/2020\|EPI_ISL_569268\|20 | ···················· | ···················· | ····················· | ···················· | ···················· | ···················· |
| K | 1712 | hCoV-19/France/PAC-IHU-0049/2020\|EPI_ISL_569269\|20 | ···················· | ···················· | ····················· | ···················· | ···················· | ···················· |
| K | 1713 | hCoV-19/France/PAC-IHU-0050/2020\|EPI_ISL_569270\|20 | ···················· | ···················· | ····················· | ···················· | ···················· | ···················· |
| K | 1714 | hCoV-19/France/PAC-IHU-0051/2020\|EPI_ISL_569271\|20 | ···················· | ···················· | ····················· | ···················· | ···················· | ···················· |
| K | 1715 | hCoV-19/France/PAC-IHU-0053/2020\|EPI_ISL_569272\|20 | ···················· | ···················· | ····················· | ···················· | ···················· | ···················· |
| K | 1716 | hCoV-19/France/PAC-IHU-0054/2020\|EPI_ISL_569273\|20 | ···················· | ···················· | ····················· | ···················· | ···················· | ···················· |
| K | 1717 | hCoV-19/France/PAC-IHU-0055/2020\|EPI_ISL_569274\|20 | ···················· | ···················· | ····················· | ···················· | ···················· | ···················· |
| K | 1718 | hCoV-19/France/PAC-IHU-0056/2020\|EPI_ISL_569275\|20 | ···················· | ···················· | ····················· | ···················· | ···················· | ···················· |
| K | 1719 | hCoV-19/France/PAC-IHU-0057/2020\|EPI_ISL_569276\|20 | ···················· | ···················· | ····················· | ···················· | ···················· | ···················· |
| K | 1720 | hCoV-19/France/PAC-IHU-0058/2020\|EPI_ISL_569277\|20 | ···················· | ···················· | ····················· | ···················· | ···················· | ···················· |
| K | 1721 | hCoV-19/France/PAC-IHU-0059/2020\|EPI_ISL_569278\|20 | ···················· | ···················· | ····················· | ···················· | ···················· | ···················· |
| K | 1722 | hCoV-19/France/PAC-IHU-0060/2020\|EPI_ISL_569279\|20 | ···················· | ···················· | ····················· | ···················· | ···················· | ···················· |
| K | 1723 | hCoV-19/France/PAC-IHU-0061/2020\|EPI_ISL_569280\|20 | ···················· | ···················· | ····················· | ···················· | ···················· | ···················· |
| K | 1725 | hCoV-19/France/PAC-IHU-0063/2020\|EPI_ISL_569282\|20 | ···················· | ···················· | ····················· | ···················· | ···················· | ···················· |
| K | 1726 | hCoV-19/France/PAC-IHU-0065/2020\|EPI_ISL_569284\|20 | ···················· | ···················· | ····················· | ···················· | ···················· | ···················· |
| K | 1727 | hCoV-19/France/PAC-IHU-0066/2020\|EPI_ISL_569285\|20 | ···················· | ···················· | ····················· | ···················· | ···················· | ···················· |
| K | 1728 | hCoV-19/France/PAC-IHU-0071/2020\|EPI_ISL_569288\|20 | ···················· | ···················· | ····················· | ···················· | ···················· | ···················· |
| K | 1729 | hCoV-19/France/PAC-IHU-0073/2020\|EPI_ISL_569290\|20 | ···················· | ···················· | ····················· | ···················· | ···················· | ···················· |
| K | 1730 | hCoV-19/France/PAC-IHU-0074/2020\|EPI_ISL_569291\|20 | ···················· | ···················· | ····················· | ···················· | ···················· | ···················· |
| K | 1731 | hCoV-19/France/PAC-IHU-0075/2020\|EPI_ISL_569292\|20 | ···················· | ···················· | ····················· | ···················· | ···················· | ···················· |
| K | 1732 | hCoV-19/France/PAC-IHU-0076/2020\|EPI_ISL_569293\|20 | ···················· | ···················· | ····················· | ···················· | ···················· | ···················· |
| K | 1733 | hCoV-19/France/PAC-IHU-0078/2020\|EPI_ISL_569294\|20 | ···················· | ···················· | ····················· | ···················· | ···················· | ···················· |
| K | 1734 | hCoV-19/France/PAC-IHU-0079/2020\|EPI_ISL_569295\|20 | ···················· | ···················· | ····················· | ···················· | ···················· | ···················· |
| K | 1735 | hCoV-19/France/PAC-IHU-0080/2020\|EPI_ISL_569296\|20 | ···················· | ···················· | ····················· | ···················· | ···················· | ···················· |
| K | 1736 | hCoV-19/France/PAC-IHU-0081/2020\|EPI_ISL_569297\|20 | ···················· | ···················· | ····················· | ···················· | ···················· | ···················· |
| K | 1737 | hCoV-19/France/PAC-IHU-0082/2020\|EPI_ISL_569298\|20 | ···················· | ···················· | ····················· | ···················· | ···················· | ···················· |
| K | 1738 | hCoV-19/France/PAC-IHU-0083/2020\|EPI_ISL_569299\|20 | ···················· | ···················· | ····················· | ···················· | ···················· | ···················· |
| K | 1739 | hCoV-19/USA/WA-UW-9301/2020\|EPI_ISL_570242\|2020-05 | ···················· | ···················· | ····················· | ···················· | ···················· | ···················· |
| K | 1740 | hCoV-19/USA/WA-UW-9308/2020\|EPI_ISL_570243\|2020-05 | ···················· | ···················· | ····················· | ···················· | ···················· | ···················· |
| K | 1741 | hCoV-19/USA/WA-UW-9333/2020\|EPI_ISL_570244\|2020-05 | ···················· | ···················· | ····················· | ···················· | ···················· | ···················· |
| K | 1742 | hCoV-19/USA/WA-UW-9338/2020\|EPI_ISL_570245\|2020-05 | ···················· | ···················· | ····················· | ···················· | ···················· | ···················· |
| K | 1743 | hCoV-19/USA/WA-UW-9339/2020\|EPI_ISL_570246\|2020-05 | ···················· | ···················· | ····················· | ···················· | ···················· | ···················· |
| K | 1745 | hCoV-19/USA/WA-UW-9361/2020\|EPI_ISL_570248\|2020-05 | ···················· | ···················· | ····················· | ···················· | ···················· | ···················· |
| K | 1746 | hCoV-19/USA/WA-UW-9362/2020\|EPI_ISL_570249\|2020-05 | ···················· | ···················· | ····················· | ···················· | ···················· | ···················· |
| K | 1747 | hCoV-19/USA/WA-UW-9374/2020\|EPI_ISL_570250\|2020-05 | ···················· | ···················· | ····················· | ···················· | ···················· | ···················· |
| K | 1748 | hCoV-19/USA/WA-UW-9407/2020\|EPI_ISL_570252\|2020-05 | ···················· | ···················· | ····················· | ···················· | ···················· | ···················· |
| K | 1749 | hCoV-19/USA/WA-UW-9422/2020\|EPI_ISL_570253\|2020-05 | ···················· | ···················· | ····················· | ···················· | ···················· | ···················· |
| K | 1750 | hCoV-19/USA/WA-UW-9439/2020\|EPI_ISL_570254\|2020-05 | ···················· | ···················· | ····················· | ···················· | ···················· | ···················· |
| K | 1751 | hCoV-19/USA/WA-UW-9500/2020\|EPI_ISL_570256\|2020-05 | ···················· | ···················· | ····················· | ···················· | ···················· | ···················· |
| K | 1752 | hCoV-19/USA/WA-UW-9504/2020\|EPI_ISL_570257\|2020-05 | ···················· | ···················· | ····················· | ···················· | ···················· | ···················· |
| K | 1753 | hCoV-19/USA/WA-UW-9520/2020\|EPI_ISL_570258\|2020-05 | ···················· | ···················· | ····················· | ···················· | ···················· | ···················· |
| K | 1754 | hCoV-19/USA/WA-UW-9532/2020\|EPI_ISL_570259\|2020-05 | ···················· | ···················· | ····················· | ···················· | ···················· | ···················· |
| K | 1755 | hCoV-19/USA/WA-UW-9534/2020\|EPI_ISL_570260\|2020-05 | ···················· | ···················· | ····················· | ···················· | ···················· | ···················· |
| K | 1756 | hCoV-19/USA/WA-UW-9536/2020\|EPI_ISL_570261\|2020-05 | ···················· | ···················· | ····················· | ···················· | ···················· | ···················· |
| K | 1757 | hCoV-19/USA/WA-UW-9568/2020\|EPI_ISL_570262\|2020-05 | ···················· | ···················· | ····················· | ···················· | ···················· | ···················· |
| K | 1758 | hCoV-19/USA/WA-UW-9578/2020\|EPI_ISL_570264\|2020-05 | ···················· | ···················· | ····················· | ···················· | ···················· | ···················· |
| K | 1759 | hCoV-19/USA/WA-UW-9579/2020\|EPI_ISL_570265\|2020-05 | ···················· | ···················· | ····················· | ···················· | ···················· | ···················· |
| K | 1760 | hCoV-19/USA/WA-UW-9580/2020\|EPI_ISL_570266\|2020-05 | ···················· | ···················· | ····················· | ···················· | ···················· | ···················· |
| K | 1761 | hCoV-19/USA/WA-UW-9600/2020\|EPI_ISL_570267\|2020-05 | ···················· | ···················· | ····················· | ···················· | ···················· | ···················· |
| K | 1762 | hCoV-19/USA/WA-UW-10110/2020\|EPI_ISL_570268\|2020-0 | ···················· | ···················· | ····················· | ···················· | ···················· | ···················· |
| K | 1763 | hCoV-19/USA/WA-UW-10158/2020\|EPI_ISL_570269\|2020-0 | ···················· | ···················· | ····················· | ···················· | ···················· | ···················· |
| K | 1764 | hCoV-19/USA/WA-UW-10159/2020\|EPI_ISL_570270\|2020-0 | ···················· | ·A·················· | ····················· | ···················· | ···················· | ···················· |
| K | 1765 | hCoV-19/USA/WA-UW-10163/2020\|EPI_ISL_570271\|2020-0 | ···················· | ···················· | ····················· | ···················· | ···················· | ···················· |
| K | 1766 | hCoV-19/USA/WA-UW-10185/2020\|EPI_ISL_570272\|2020-0 | ···················· | ···················· | ····················· | ···················· | ···················· | ···················· |
| K | 1767 | hCoV-19/USA/WA-UW-10187/2020\|EPI_ISL_570273\|2020-0 | ···················· | ···················· | ····················· | ···················· | ···················· | ···················· |
| K | 1768 | hCoV-19/USA/WA-UW-10189/2020\|EPI_ISL_570274\|2020-0 | ···················· | ···················· | ····················· | ···················· | ···················· | ···················· |

| | | | | | | | | |
|---|---|---|---|---|---|---|---|---|
| K | 1769 | hCoV-19/USA/WA-UW-10190/2020\|EPI_ISL_570275\|2020-0 | ···················· | ···················· | ····················· | ···················· | ···················· | ···················· |
| K | 1770 | hCoV-19/USA/WA-UW-10191/2020\|EPI_ISL_570276\|2020-0 | ···················· | ···················· | ····················· | ···················· | ···················· | ···················· |
| K | 1771 | hCoV-19/USA/WA-UW-10197/2020\|EPI_ISL_570277\|2020-0 | ···················· | ···················· | ····················· | ···················· | ···················· | ···················· |
| K | 1772 | hCoV-19/USA/WA-UW-10199/2020\|EPI_ISL_570278\|2020-0 | ···················· | ···················· | ····················· | ···················· | ···················· | ···················· |
| K | 1773 | hCoV-19/USA/WA-UW-10206/2020\|EPI_ISL_570279\|2020-0 | ···················· | ···················· | ····················· | ···················· | ···················· | ···················· |
| K | 1774 | hCoV-19/USA/WA-UW-10213/2020\|EPI_ISL_570280\|2020-0 | ···················· | ···················· | ····················· | ···················· | ···················· | ···················· |
| K | 1775 | hCoV-19/USA/WA-UW-10216/2020\|EPI_ISL_570281\|2020-0 | ···················· | ···················· | ····················· | ···················· | ···················· | ···················· |
| K | 1776 | hCoV-19/USA/WA-UW-10217/2020\|EPI_ISL_570282\|2020-0 | ···················· | ···················· | ····················· | ···················· | ···················· | ···················· |
| K | 1777 | hCoV-19/USA/WA-UW-10230/2020\|EPI_ISL_570283\|2020-0 | ···················· | ···················· | ····················· | ···················· | ···················· | ···················· |
| K | 1778 | hCoV-19/USA/WA-UW-10231/2020\|EPI_ISL_570284\|2020-0 | ···················· | ···················· | ····················· | ···················· | ···················· | ···················· |
| K | 1779 | hCoV-19/USA/WA-UW-10247/2020\|EPI_ISL_570285\|2020-0 | ···················· | ···················· | ····················· | ···················· | ···················· | ···················· |
| K | 1780 | hCoV-19/USA/WA-UW-10250/2020\|EPI_ISL_570286\|2020-0 | ···················· | ···················· | ····················· | ···················· | ···················· | ···················· |
| K | 1781 | hCoV-19/USA/WA-UW-10272/2020\|EPI_ISL_570287\|2020-0 | ···················· | ···················· | ····················· | ···················· | ···················· | ···················· |
| K | 1782 | hCoV-19/USA/WA-UW-10277/2020\|EPI_ISL_570288\|2020-0 | ···················· | ···················· | ····················· | ···················· | ···················· | ···················· |
| K | 1783 | hCoV-19/USA/WA-UW-10283/2020\|EPI_ISL_570289\|2020-0 | ···················· | ···················· | ····················· | ···················· | ···················· | ···················· |
| K | 1784 | hCoV-19/USA/WA-UW-10284/2020\|EPI_ISL_570290\|2020-0 | ···················· | ···················· | ····················· | ···················· | ···················· | ···················· |
| K | 1785 | hCoV-19/USA/WA-UW-10286/2020\|EPI_ISL_570291\|2020-0 | ···················· | ···················· | ····················· | ···················· | ···················· | ···················· |
| K | 1786 | hCoV-19/USA/WA-UW-10298/2020\|EPI_ISL_570292\|2020-0 | ···················· | ···················· | ····················· | ···················· | ···················· | ···················· |
| K | 1787 | hCoV-19/USA/WA-UW-10305/2020\|EPI_ISL_570293\|2020-0 | ···················· | ···················· | ····················· | ···················· | ···················· | ···················· |
| K | 1788 | hCoV-19/USA/WA-UW-10306/2020\|EPI_ISL_570294\|2020-0 | ···················· | ···················· | ····················· | ···················· | ···················· | ···················· |
| K | 1789 | hCoV-19/USA/WA-UW-10317/2020\|EPI_ISL_570295\|2020-0 | ···················· | ···················· | ····················· | ···················· | ···················· | ···················· |
| K | 1790 | hCoV-19/USA/WA-UW-10365/2020\|EPI_ISL_570297\|2020-0 | ···················· | ···················· | ····················· | ···················· | ···················· | ···················· |
| K | 1791 | hCoV-19/USA/WA-UW-10380/2020\|EPI_ISL_570298\|2020-0 | ···················· | ···················· | ····················· | ···················· | ···················· | ···················· |
| K | 1792 | hCoV-19/USA/WA-UW-10392/2020\|EPI_ISL_570299\|2020-0 | ···················· | ···················· | ····················· | ···················· | ···················· | ···················· |
| K | 1793 | hCoV-19/England/GSTT-5658/2020\|EPI_ISL_560700\|2020 | ···················· | ···················· | ····················· | ···················· | ···················· | ···················· |
| K | 1794 | hCoV-19/England/GSTT-5659/2020\|EPI_ISL_560701\|2020 | ···················· | ···················· | ····················· | ···················· | ···················· | ···················· |
| K | 1795 | hCoV-19/England/GSTT-5688/2020\|EPI_ISL_560702\|2020 | ···················· | ···················· | ····················· | ···················· | ···················· | ···················· |
| K | 1796 | hCoV-19/England/GSTT-5705/2020\|EPI_ISL_560705\|2020 | ···················· | ···················· | ····················· | ···················· | ···················· | ···················· |
| K | 1797 | hCoV-19/England/GSTT-5777/2020\|EPI_ISL_560712\|2020 | ···················· | ···················· | ····················· | ···················· | ···················· | ···················· |
| K | 1798 | hCoV-19/England/GSTT-6706/2020\|EPI_ISL_560719\|2020 | ···················· | ···················· | ····················· | ···················· | ···················· | ···················· |
| K | 1799 | hCoV-19/USA/MN-MDH-1683/2020\|EPI_ISL_560770\|2020-0 | ···················· | ···················· | ····················· | ···················· | ···················· | ···················· |
| K | 1800 | hCoV-19/USA/MN-MDH-1695/2020\|EPI_ISL_560782\|2020-0 | ···················· | ···················· | ····················· | ···················· | ···················· | ···················· |
| K | 1801 | hCoV-19/USA/MN-MDH-1696/2020\|EPI_ISL_560783\|2020-0 | ···················· | ···················· | ····················· | ···················· | ···················· | ···················· |
| K | 1802 | hCoV-19/USA/MN-MDH-1697/2020\|EPI_ISL_560784\|2020-0 | ···················· | ···················· | ····················· | ···················· | ···················· | ···················· |
| K | 1803 | hCoV-19/USA/MN-MDH-1698/2020\|EPI_ISL_560785\|2020-0 | ···················· | ···················· | ····················· | ···················· | ···················· | ···················· |
| K | 1804 | hCoV-19/USA/MN-MDH-1699/2020\|EPI_ISL_560786\|2020-0 | ···················· | ···················· | ····················· | ···················· | ···················· | ···················· |
| K | 1805 | hCoV-19/USA/MN-MDH-1700/2020\|EPI_ISL_560787\|2020-0 | ···················· | ···················· | ····················· | ···················· | ···················· | ···················· |
| K | 1806 | hCoV-19/USA/MN-MDH-1701/2020\|EPI_ISL_560788\|2020-0 | ···················· | ···················· | ····················· | ···················· | ···················· | ···················· |
| K | 1807 | hCoV-19/USA/MN-MDH-1702/2020\|EPI_ISL_560789\|2020-0 | ···················· | ···················· | ····················· | ···················· | ···················· | ···················· |
| K | 1808 | hCoV-19/USA/MN-MDH-1703/2020\|EPI_ISL_560790\|2020-0 | ···················· | ···G················ | ····················· | ···················· | ···················· | ···················· |
| K | 1809 | hCoV-19/USA/MN-MDH-1704/2020\|EPI_ISL_560791\|2020-0 | ···················· | ···················· | ····················· | ···················· | ···················· | ···················· |
| K | 1810 | hCoV-19/USA/MN-MDH-1705/2020\|EPI_ISL_560792\|2020-0 | ········T··········· | ···················· | ····················· | ···················· | ···················· | ···················· |
| K | 1811 | hCoV-19/USA/MN-MDH-1706/2020\|EPI_ISL_560793\|2020-0 | ···················· | ···················· | ····················· | ···················· | ···················· | ···················· |
| K | 1812 | hCoV-19/USA/MN-MDH-1707/2020\|EPI_ISL_560794\|2020-0 | ···················· | ···················· | ····················· | ···················· | ···················· | ···················· |
| K | 1813 | hCoV-19/USA/MN-MDH-1708/2020\|EPI_ISL_560795\|2020-0 | ···················· | ···················· | ····················· | ···················· | ···················· | ···················· |
| K | 1814 | hCoV-19/USA/MN-MDH-1709/2020\|EPI_ISL_560796\|2020-0 | ···················· | ···················· | ····················· | ···················· | ···················· | ···················· |
| K | 1815 | hCoV-19/USA/MN-MDH-1710/2020\|EPI_ISL_560797\|2020-0 | ···················· | ···················· | ····················· | ···················· | ···················· | ···················· |
| K | 1816 | hCoV-19/USA/MN-MDH-1711/2020\|EPI_ISL_560798\|2020-0 | ···················· | ···················· | ····················· | ···················· | ···················· | ···················· |
| K | 1817 | hCoV-19/USA/MN-MDH-1712/2020\|EPI_ISL_560799\|2020-0 | ···················· | ···················· | ····················· | ···················· | ···················· | ···················· |
| K | 1818 | hCoV-19/Australia/VIC10641/2020\|EPI_ISL_563008\|202 | ···················· | ···················· | ····················· | ···················· | ···················· | ···················· |
| K | 1819 | hCoV-19/Australia/VIC10642/2020\|EPI_ISL_563009\|202 | ···················· | ···················· | ····················· | ···················· | ···················· | ···················· |
| K | 1820 | hCoV-19/Australia/VIC7128/2020\|EPI_ISL_564247\|2020 | ···················· | ···················· | ····················· | ···················· | ···················· | ···················· |
| K | 1821 | hCoV-19/Australia/VIC7129/2020\|EPI_ISL_564248\|2020 | ···················· | ···················· | ····················· | ···················· | ···················· | ···················· |
| K | 1822 | hCoV-19/Australia/VIC7132/2020\|EPI_ISL_564249\|2020 | ···················· | ···················· | ····················· | ···················· | ···················· | ···················· |
| K | 1823 | hCoV-19/Australia/VIC7135/2020\|EPI_ISL_564250\|2020 | ···················· | ···················· | ····················· | ···················· | ···················· | ···················· |
| K | 1824 | hCoV-19/Australia/VIC7136/2020\|EPI_ISL_564251\|2020 | ···················· | ···················· | ····················· | ···················· | ···················· | ···················· |
| K | 1825 | hCoV-19/Australia/VIC7139/2020\|EPI_ISL_564252\|2020 | ···················· | ···················· | ···K················· | ···················· | ···················· | ···················· |
| K | 1826 | hCoV-19/Australia/VIC7145/2020\|EPI_ISL_564253\|2020 | ···················· | ···················· | ····················· | ···················· | ···················· | ···················· |
| K | 1827 | hCoV-19/Australia/VIC7147/2020\|EPI_ISL_564254\|2020 | ···················· | ···················· | ····················· | ···················· | ···················· | ···················· |
| K | 1828 | hCoV-19/Australia/VIC7158/2020\|EPI_ISL_564255\|2020 | ···················· | ···················· | ····················· | ···················· | ···················· | ···················· |
| K | 1829 | hCoV-19/Australia/VIC7162/2020\|EPI_ISL_564256\|2020 | ···················· | ···················· | ····················· | ···················· | ···S················ | ···················· |
| K | 1830 | hCoV-19/Australia/VIC7163/2020\|EPI_ISL_564257\|2020 | ···················· | ···················· | ····················· | ···················· | ···················· | ···················· |
| K | 1831 | hCoV-19/Australia/VIC7164/2020\|EPI_ISL_564258\|2020 | ···················· | ···················· | ····················· | ···················· | ···················· | ···················· |
| K | 1832 | hCoV-19/Australia/VIC7165/2020\|EPI_ISL_564259\|2020 | ···················· | ···················· | ····················· | ···················· | ···················· | ···················· |
| K | 1833 | hCoV-19/Australia/VIC7170/2020\|EPI_ISL_564260\|2020 | ···················· | ···················· | ····················· | ···················· | ···················· | ···················· |
| K | 1834 | hCoV-19/Australia/VIC7173/2020\|EPI_ISL_564261\|2020 | ···················· | ···················· | ····················· | ···················· | ···················· | ···················· |
| K | 1835 | hCoV-19/Australia/VIC7175/2020\|EPI_ISL_564262\|2020 | ···················· | ···················· | ····················· | ···················· | ···················· | ···················· |
| K | 1836 | hCoV-19/Australia/VIC7176/2020\|EPI_ISL_564263\|2020 | ···················· | ···················· | ····················· | ···················· | ···················· | ···················· |
| K | 1837 | hCoV-19/Australia/VIC7177/2020\|EPI_ISL_564264\|2020 | ···················· | ···················· | ····················· | ···················· | ···················· | ···················· |
| K | 1838 | hCoV-19/Australia/VIC7178/2020\|EPI_ISL_564265\|2020 | ···················· | ···················· | ····················· | ···················· | ···················· | ···················· |
| K | 1839 | hCoV-19/Australia/VIC7179/2020\|EPI_ISL_564266\|2020 | ···················· | ···················· | ····················· | ···················· | ···················· | ···················· |
| K | 1840 | hCoV-19/Australia/VIC7181/2020\|EPI_ISL_564267\|2020 | ···················· | ···················· | ····················· | ···················· | ···················· | ···················· |
| K | 1841 | hCoV-19/Australia/VIC7185/2020\|EPI_ISL_564268\|2020 | ···················· | ···················· | ····················· | ···················· | ···················· | ···················· |
| K | 1842 | hCoV-19/Australia/VIC7189/2020\|EPI_ISL_564269\|2020 | ···················· | ···················· | ····················· | ···················· | ···················· | ···················· |
| K | 1844 | hCoV-19/Australia/VIC7190/2020\|EPI_ISL_564271\|2020 | ···················· | ···················· | ····················· | ···················· | ···················· | ···················· |
| K | 1845 | hCoV-19/Australia/VIC7191/2020\|EPI_ISL_564272\|2020 | ···················· | ···················· | ····················· | ···················· | ···················· | ···················· |
| K | 1846 | hCoV-19/Australia/VIC7192/2020\|EPI_ISL_564273\|2020 | ···················· | ···················· | ····················· | ···················· | ···················· | ···················· |
| K | 1847 | hCoV-19/Australia/VIC7193/2020\|EPI_ISL_564274\|2020 | ···················· | ···················· | ····················· | ···················· | ···················· | ···················· |
| K | 1848 | hCoV-19/Australia/VIC7195/2020\|EPI_ISL_564275\|2020 | ···················· | ···················· | ····················· | ···················· | ···················· | ···················· |
| K | 1849 | hCoV-19/Australia/VIC7196/2020\|EPI_ISL_564276\|2020 | ···················· | ···················· | ····················· | ···················· | ···················· | ···················· |

| | | | | | | | | |
|---|---|---|---|---|---|---|---|---|
| K | 1850 | hCoV-19/Australia/VIC7199/2020\|EPI_ISL_564277\|2020 | ···················· | ···················· | ····················· | ···················· | ···················· | ···················· |
| K | 1851 | hCoV-19/Australia/VIC7201/2020\|EPI_ISL_564278\|2020 | ···················· | ···················· | ····················· | ···················· | ···················· | ···················· |
| K | 1852 | hCoV-19/Australia/VIC7202/2020\|EPI_ISL_564279\|2020 | ···················· | ···················· | ····················· | ···················· | ···················· | ···················· |
| K | 1853 | hCoV-19/Australia/VIC7203/2020\|EPI_ISL_564280\|2020 | ···················· | ···················· | ····················· | ···················· | ···················· | ···················· |
| K | 1854 | hCoV-19/Australia/VIC7206/2020\|EPI_ISL_564281\|2020 | ···················· | ···················· | ····················· | ···················· | ···················· | ···················· |
| K | 1855 | hCoV-19/Australia/VIC7207/2020\|EPI_ISL_564282\|2020 | ···················· | ···················· | ····················· | ···················· | ···················· | ···················· |
| K | 1856 | hCoV-19/Australia/VIC7208/2020\|EPI_ISL_564283\|2020 | ···················· | ···················· | ····················· | ···················· | ···················· | ···················· |
| K | 1857 | hCoV-19/Australia/VIC7209/2020\|EPI_ISL_564284\|2020 | ···················· | ···················· | ····················· | ···················· | ···················· | ···················· |
| K | 1858 | hCoV-19/Australia/VIC7210/2020\|EPI_ISL_564285\|2020 | ···················· | ···················· | ····················· | ···················· | ···················· | ···················· |
| K | 1859 | hCoV-19/Australia/VIC7212/2020\|EPI_ISL_564286\|2020 | ···················· | ···················· | ····················· | ···················· | ···················· | ···················· |
| K | 1860 | hCoV-19/Australia/VIC7214/2020\|EPI_ISL_564287\|2020 | ···················· | ···················· | ····················· | ···················· | ···················· | ···················· |
| K | 1861 | hCoV-19/Australia/VIC7215/2020\|EPI_ISL_564288\|2020 | ···················· | ···················· | ····················· | ···················· | ···················· | ···················· |
| K | 1862 | hCoV-19/Australia/VIC7216/2020\|EPI_ISL_564289\|2020 | ···················· | ···················· | ····················· | ···················· | ···················· | ···················· |
| K | 1863 | hCoV-19/Australia/VIC7219/2020\|EPI_ISL_564290\|2020 | ···················· | ···················· | ····················· | ···················· | ···················· | ···················· |
| K | 1864 | hCoV-19/Australia/VIC7220/2020\|EPI_ISL_564291\|2020 | ···················· | ···················· | ····················· | ···················· | ···················· | ···················· |
| K | 1865 | hCoV-19/Australia/VIC7221/2020\|EPI_ISL_564292\|2020 | ···················· | ···················· | ····················· | ···················· | ···················· | ···················· |
| K | 1866 | hCoV-19/Australia/VIC7223/2020\|EPI_ISL_564293\|2020 | ···················· | ···················· | ····················· | ···················· | ···················· | ···················· |
| K | 1867 | hCoV-19/Australia/VIC7226/2020\|EPI_ISL_564294\|2020 | ···················· | ···················· | ····················· | ···················· | ···················· | ···················· |
| K | 1868 | hCoV-19/Australia/VIC7227/2020\|EPI_ISL_564295\|2020 | ···················· | ···················· | ····················· | ···················· | ···················· | ···················· |
| K | 1869 | hCoV-19/Australia/VIC7229/2020\|EPI_ISL_564296\|2020 | ···················· | ···················· | ····················· | ···················· | ···················· | ···················· |
| K | 1870 | hCoV-19/Australia/VIC7230/2020\|EPI_ISL_564297\|2020 | ···················· | ···················· | ····················· | ···················· | ···················· | ···················· |
| K | 1871 | hCoV-19/Australia/VIC7231/2020\|EPI_ISL_564298\|2020 | ···················· | ···················· | ····················· | ···················· | ···················· | ···················· |
| K | 1872 | hCoV-19/Australia/VIC7232/2020\|EPI_ISL_564299\|2020 | ···················· | ···················· | ····················· | ···················· | ···················· | ···················· |
| K | 1873 | hCoV-19/Australia/VIC7233/2020\|EPI_ISL_564300\|2020 | ···················· | ·········A·········· | ····················· | ···················· | ···················· | ···················· |
| K | 1874 | hCoV-19/Australia/VIC7234/2020\|EPI_ISL_564301\|2020 | ···················· | ···················· | ····················· | ···················· | ···················· | ···················· |
| K | 1875 | hCoV-19/Australia/VIC7235/2020\|EPI_ISL_564302\|2020 | ···················· | ···················· | ····················· | ···················· | ···················· | ···················· |
| K | 1876 | hCoV-19/Australia/VIC7236/2020\|EPI_ISL_564303\|2020 | ···················· | ···················· | ····················· | ···················· | ···················· | ···················· |
| K | 1877 | hCoV-19/Australia/VIC7237/2020\|EPI_ISL_564304\|2020 | ···················· | ···················· | ····················· | ···················· | ···················· | ···················· |
| K | 1878 | hCoV-19/Australia/VIC7238/2020\|EPI_ISL_564305\|2020 | ···················· | ···················· | ····················· | ···················· | ···················· | ···················· |
| K | 1879 | hCoV-19/Australia/VIC7239/2020\|EPI_ISL_564306\|2020 | ···················· | ···················· | ····················· | ···················· | ···················· | ···················· |
| K | 1880 | hCoV-19/Australia/VIC7241/2020\|EPI_ISL_564307\|2020 | ···················· | ···················· | ····················· | ···················· | ···················· | ···················· |
| K | 1881 | hCoV-19/Australia/VIC7242/2020\|EPI_ISL_564308\|2020 | ···················· | ···················· | ····················· | ···················· | ···················· | ···················· |
| K | 1882 | hCoV-19/Australia/VIC7243/2020\|EPI_ISL_564309\|2020 | ···················· | ···················· | ····················· | ···················· | ···················· | ···················· |
| K | 1883 | hCoV-19/Australia/VIC7244/2020\|EPI_ISL_564310\|2020 | ···················· | ···················· | ····················· | ···················· | ···················· | ···················· |
| K | 1884 | hCoV-19/Australia/VIC7248/2020\|EPI_ISL_564311\|2020 | ···················· | ···················· | ····················· | ···················· | ···················· | ···················· |
| K | 1885 | hCoV-19/Australia/VIC7249/2020\|EPI_ISL_564312\|2020 | ···················· | ···················· | ····················· | ···················· | ···················· | ···················· |
| K | 1886 | hCoV-19/Australia/VIC7250/2020\|EPI_ISL_564313\|2020 | ···················· | ···················· | ····················· | ···················· | ···················· | ···················· |
| K | 1887 | hCoV-19/Australia/VIC7251/2020\|EPI_ISL_564314\|2020 | ···················· | ···················· | ····················· | ···················· | ···················· | ···················· |
| K | 1888 | hCoV-19/Australia/VIC7252/2020\|EPI_ISL_564315\|2020 | ···················· | ···················· | ····················· | ···················· | ···················· | ···················· |
| K | 1889 | hCoV-19/Australia/VIC7253/2020\|EPI_ISL_564316\|2020 | ···················· | ···················· | ····················· | ···················· | ···················· | ···················· |
| K | 1890 | hCoV-19/Australia/VIC7254/2020\|EPI_ISL_564317\|2020 | ···················· | ···················· | ····················· | ···················· | ···················· | ···················· |
| K | 1891 | hCoV-19/Australia/VIC7257/2020\|EPI_ISL_564318\|2020 | ···················· | ···················· | ····················· | ···················· | ···················· | ···················· |
| K | 1892 | hCoV-19/Australia/VIC7260/2020\|EPI_ISL_564319\|2020 | ···················· | ···················· | ····················· | ···················· | ···················· | ···················· |
| K | 1893 | hCoV-19/Australia/VIC7263/2020\|EPI_ISL_564320\|2020 | ···················· | ···················· | ····················· | ···················· | ···················· | ···················· |
| K | 1894 | hCoV-19/Australia/VIC7264/2020\|EPI_ISL_564321\|2020 | ···················· | ···················· | ····················· | ···················· | ···················· | ···················· |
| K | 1895 | hCoV-19/Australia/VIC7268/2020\|EPI_ISL_564322\|2020 | ···················· | ···················· | ····················· | ···················· | ···················· | ···················· |
| K | 1896 | hCoV-19/Australia/VIC7271/2020\|EPI_ISL_564323\|2020 | ···················· | ···················· | ··C·················· | ···················· | ···················· | ···················· |
| K | 1897 | hCoV-19/Australia/VIC7311/2020\|EPI_ISL_564339\|2020 | ···················· | ···················· | ····················· | ···················· | ······Y············· | ···················· |
| K | 1898 | hCoV-19/Australia/VIC9505/2020\|EPI_ISL_565578\|2020 | ···················· | ···················· | ····················· | ···················· | ···················· | ···················· |
| K | 1899 | hCoV-19/Australia/VIC9506/2020\|EPI_ISL_565579\|2020 | ···················· | ···················· | ····················· | ···················· | ···················· | ···················· |
| K | 1900 | hCoV-19/Australia/VIC9508/2020\|EPI_ISL_565580\|2020 | ···················· | ···················· | ··S·················· | ···················· | ···················· | ···················· |
| K | 1901 | hCoV-19/Australia/VIC9512/2020\|EPI_ISL_565581\|2020 | ···················· | ···················· | ····················· | ···················· | ···················· | ···················· |
| K | 1902 | hCoV-19/Australia/VIC9516/2020\|EPI_ISL_565582\|2020 | ···················· | ···················· | ····················· | ···················· | ···················· | ···················· |
| K | 1904 | hCoV-19/Australia/VIC9520/2020\|EPI_ISL_565584\|2020 | ···················· | ···················· | ····················· | ···················· | ···················· | ···················· |
| K | 1905 | hCoV-19/Australia/VIC9521/2020\|EPI_ISL_565585\|2020 | ···················· | ···················· | ····················· | ···················· | ···················· | ···················· |
| K | 1906 | hCoV-19/Australia/VIC9522/2020\|EPI_ISL_565586\|2020 | ···················· | ···················· | ····················· | ···················· | ···················· | ···················· |
| K | 1907 | hCoV-19/Australia/VIC9524/2020\|EPI_ISL_565587\|2020 | ···················· | ···················· | ····················· | ···················· | ···················· | ···················· |
| K | 1908 | hCoV-19/Australia/VIC9526/2020\|EPI_ISL_565588\|2020 | ···················· | ···················· | ····················· | ···················· | ···················· | ···················· |
| K | 1909 | hCoV-19/Australia/VIC9528/2020\|EPI_ISL_565589\|2020 | ···················· | ···················· | ····················· | ···················· | ···················· | ···················· |
| K | 1910 | hCoV-19/Australia/VIC9533/2020\|EPI_ISL_565590\|2020 | ···················· | ···················· | ····················· | ···················· | ···················· | ···················· |
| K | 1911 | hCoV-19/Australia/VIC9534/2020\|EPI_ISL_565591\|2020 | ···················· | ···················· | ····················· | ···················· | ···················· | ···················· |
| K | 1912 | hCoV-19/Australia/VIC9537/2020\|EPI_ISL_565592\|2020 | ···················· | ···················· | ····················· | ···················· | ···················· | ···················· |
| K | 1914 | hCoV-19/Australia/VIC9541/2020\|EPI_ISL_565594\|2020 | ···················· | ···················· | ····················· | ···················· | ···················· | ···················· |
| K | 1915 | hCoV-19/Australia/VIC9543/2020\|EPI_ISL_565595\|2020 | ···················· | ···················· | ····················· | ···················· | ···················· | ···················· |
| K | 1916 | hCoV-19/Australia/VIC9544/2020\|EPI_ISL_565596\|2020 | ···················· | ···················· | ····················· | ···················· | ···················· | ···················· |
| K | 1917 | hCoV-19/Australia/VIC9545/2020\|EPI_ISL_565597\|2020 | ···················· | ···················· | ····················· | ···················· | ···················· | ···················· |
| K | 1918 | hCoV-19/Australia/VIC9551/2020\|EPI_ISL_565598\|2020 | ···················· | ···················· | ····················· | ···················· | ···················· | ···················· |
| K | 1919 | hCoV-19/Australia/VIC9555/2020\|EPI_ISL_565599\|2020 | ···················· | ···················· | ····················· | ···················· | ···················· | ···················· |
| K | 1920 | hCoV-19/Australia/VIC9582/2020\|EPI_ISL_565611\|2020 | ···················· | ···················· | ····················· | ···················· | ···················· | ···················· |
| K | 1921 | hCoV-19/Australia/VIC9583/2020\|EPI_ISL_565612\|2020 | ···················· | ···················· | ····················· | ···················· | ···················· | ···················· |
| K | 1922 | hCoV-19/Australia/VIC9584/2020\|EPI_ISL_565613\|2020 | ···················· | ···················· | ····················· | ···················· | ···················· | ···················· |
| K | 1923 | hCoV-19/Australia/VIC9585/2020\|EPI_ISL_565614\|2020 | ···················· | ···················· | ····················· | ···················· | ···················· | ···················· |
| K | 1924 | hCoV-19/Australia/VIC9586/2020\|EPI_ISL_565615\|2020 | ···················· | ···················· | ····················· | ···················· | ···················· | ···················· |
| K | 1925 | hCoV-19/Australia/VIC9587/2020\|EPI_ISL_565616\|2020 | ···················· | ···················· | ····················· | ···················· | ···················· | ···················· |
| K | 1926 | hCoV-19/Australia/VIC9595/2020\|EPI_ISL_565617\|2020 | ···················· | ···················· | ····················· | ···················· | ···················· | ···················· |
| K | 1927 | hCoV-19/Australia/VIC9597/2020\|EPI_ISL_565618\|2020 | ···················· | ···················· | ····················· | ···················· | ···················· | ···················· |
| K | 1928 | hCoV-19/Australia/VIC9599/2020\|EPI_ISL_565619\|2020 | ···················· | ···················· | ····················· | ···················· | ···················· | ···················· |
| K | 1929 | hCoV-19/Australia/VIC9600/2020\|EPI_ISL_565620\|2020 | ···················· | ···················· | ····················· | ···················· | ···················· | ···················· |
| K | 1930 | hCoV-19/Australia/VIC9603/2020\|EPI_ISL_565621\|2020 | ···················· | ···················· | ····················· | ···················· | ···················· | ···················· |
| K | 1931 | hCoV-19/Australia/VIC9604/2020\|EPI_ISL_565622\|2020 | ···················· | ···················· | ····················· | ···················· | ···················· | ···················· |

| | | | | | | | | | |
|---|---|---|---|---|---|---|---|---|---|
| K | 1932 | hCoV-19/Australia/VIC9609/2020\|EPI_ISL_565623\|2020 | ···················· | ···················· | ···················· | ···················· | ···················· | ···················· | |
| K | 1933 | hCoV-19/Australia/VIC9610/2020\|EPI_ISL_565624\|2020 | ···················· | ···················· | ···················· | ···················· | NNN················· | ···················· | 1 |
| K | 1937 | hCoV-19/Australia/VIC9618/2020\|EPI_ISL_565628\|2020 | ···················· | ···················· | ···················· | ···················· | ···················· | ···················· | |
| K | 1941 | hCoV-19/Australia/VIC9625/2020\|EPI_ISL_565632\|2020 | ···················· | ···················· | ···················· | ···················· | ···················· | ···················· | |
| K | 1942 | hCoV-19/Australia/VIC9638/2020\|EPI_ISL_565633\|2020 | ···················· | ···················· | ···················· | ···················· | ···················· | ···················· | |
| K | 1943 | hCoV-19/Australia/VIC9640/2020\|EPI_ISL_565634\|2020 | ···················· | ···················· | ···················· | ···················· | ···················· | ···················· | |
| K | 1944 | hCoV-19/Australia/VIC9642/2020\|EPI_ISL_565635\|2020 | ···················· | ···················· | ···················· | ···················· | ···················· | ···················· | |
| K | 1945 | hCoV-19/Australia/VIC9645/2020\|EPI_ISL_565636\|2020 | ···················· | ···················· | ···················· | ···················· | ···················· | ···················· | |
| K | 1946 | hCoV-19/Australia/VIC9646/2020\|EPI_ISL_565637\|2020 | ···················· | ···················· | ···················· | ···················· | ···················· | ···················· | |
| K | 1947 | hCoV-19/Australia/VIC9651/2020\|EPI_ISL_565638\|2020 | ···················· | ···················· | ···················· | ···················· | ···················· | ···················· | |
| K | 1948 | hCoV-19/Australia/VIC9652/2020\|EPI_ISL_565639\|2020 | ···················· | ···················· | ···················· | ···················· | ···················· | ···················· | |
| K | 1949 | hCoV-19/Australia/VIC9654/2020\|EPI_ISL_565640\|2020 | ···················· | ···················· | ···················· | ···················· | ···················· | ···················· | |
| K | 1950 | hCoV-19/Australia/VIC9662/2020\|EPI_ISL_565641\|2020 | ···················· | ···················· | ···················· | ···················· | ···················· | ···················· | |
| K | 1951 | hCoV-19/Australia/VIC9664/2020\|EPI_ISL_565642\|2020 | ···················· | ···················· | ···················· | ···················· | ···················· | ···················· | |
| K | 1952 | hCoV-19/Australia/VIC9666/2020\|EPI_ISL_565643\|2020 | ···················· | ···················· | ···················· | ···················· | ···················· | ···················· | |
| K | 1953 | hCoV-19/Australia/VIC9667/2020\|EPI_ISL_565644\|2020 | ···················· | ···················· | ···················· | ···················· | ···················· | ···················· | |
| K | 1954 | hCoV-19/Australia/VIC9668/2020\|EPI_ISL_565645\|2020 | ···················· | ···················· | ···················· | ···················· | ···················· | ···················· | |
| K | 1955 | hCoV-19/Australia/VIC9673/2020\|EPI_ISL_565646\|2020 | ···················· | ···················· | ···················· | ···················· | ···················· | ···················· | |
| K | 1956 | hCoV-19/Australia/VIC9675/2020\|EPI_ISL_565647\|2020 | ···················· | ···················· | ···················· | ···················· | ···················· | ···················· | |
| K | 1957 | hCoV-19/Australia/VIC9676/2020\|EPI_ISL_565648\|2020 | ···················· | ···················· | ···················· | ···················· | ···················· | ···················· | |
| K | 1958 | hCoV-19/Australia/VIC9677/2020\|EPI_ISL_565649\|2020 | ···················· | ···················· | ···················· | ···················· | ···················· | ···················· | |
| K | 1959 | hCoV-19/Australia/VIC9680/2020\|EPI_ISL_565650\|2020 | ···················· | ···················· | ···················· | ···················· | ···················· | ···················· | |
| K | 1960 | hCoV-19/Australia/VIC9682/2020\|EPI_ISL_565651\|2020 | ···················· | ···················· | ···················· | ···················· | ···················· | ···················· | |
| K | 1961 | hCoV-19/Australia/VIC9685/2020\|EPI_ISL_565652\|2020 | ···················· | ···················· | ···················· | ···················· | ···················· | ···················· | |
| K | 1962 | hCoV-19/Australia/VIC9686/2020\|EPI_ISL_565653\|2020 | ···················· | ···················· | ···················· | ···················· | ···················· | ···················· | |
| K | 1963 | hCoV-19/Australia/VIC9688/2020\|EPI_ISL_565654\|2020 | ···················· | ···················· | ···················· | ···················· | ···················· | ···················· | |
| K | 1964 | hCoV-19/England/MILK-9C320A/2020\|EPI_ISL_566942\|20 | ···················· | ···················· | ···················· | ···················· | ···················· | ···················· | |
| K | 1965 | hCoV-19/England/MILK-9C3E51/2020\|EPI_ISL_566943\|20 | ···················· | ···················· | ···················· | ···················· | ···················· | ···················· | |
| K | 1966 | hCoV-19/England/MILK-9C4087/2020\|EPI_ISL_566944\|20 | ···················· | ···················· | ···················· | ···················· | ···················· | ···················· | |
| K | 1967 | hCoV-19/England/MILK-9C3862/2020\|EPI_ISL_566945\|20 | ···················· | ···················· | ···················· | ···················· | ···················· | ···················· | |
| K | 1968 | hCoV-19/England/MILK-9C3CEE/2020\|EPI_ISL_566946\|20 | ···················· | ···················· | ···················· | ···················· | ···················· | ···················· | |
| K | 1969 | hCoV-19/Wales/MILK-9C3923/2020\|EPI_ISL_566947\|2020 | ···················· | ···················· | ···················· | ···················· | ···················· | ···················· | |
| K | 1970 | hCoV-19/England/MILK-9C405A/2020\|EPI_ISL_566948\|20 | ···················· | ···················· | ···················· | ···················· | ···················· | ···················· | |
| K | 1971 | hCoV-19/England/MILK-9C3CB1/2020\|EPI_ISL_566949\|20 | ···················· | ···················· | ···················· | ···················· | ···················· | ···················· | |
| K | 1972 | hCoV-19/England/MILK-9C39AB/2020\|EPI_ISL_566950\|20 | ···················· | ·A·················· | ···················· | ···················· | ···················· | ···A················ | |
| K | 1973 | hCoV-19/England/MILK-9C43E8/2020\|EPI_ISL_566952\|20 | ···················· | ····G··············· | ···················· | ···················· | ···················· | ···················· | |
| K | 1974 | hCoV-19/England/MILK-9C35A7/2020\|EPI_ISL_566953\|20 | ···················· | ···················· | ···················· | ···················· | ···················· | ···················· | |
| K | 1975 | hCoV-19/England/MILK-9C404B/2020\|EPI_ISL_566954\|20 | ···················· | ···················· | ···················· | ···················· | ···················· | ···················· | |
| K | 1976 | hCoV-19/England/MILK-9C3ECA/2020\|EPI_ISL_566955\|20 | ···················· | ···················· | ···················· | ···················· | ···················· | ···················· | |
| K | 1977 | hCoV-19/England/MILK-9C463A/2020\|EPI_ISL_566956\|20 | ···················· | ···················· | ···················· | ···················· | ···················· | ···················· | |
| K | 1978 | hCoV-19/Wales/MILK-9C37DE/2020\|EPI_ISL_566957\|2020 | ···················· | ···················· | ···················· | ···················· | ···················· | ···················· | |
| K | 1979 | hCoV-19/England/MILK-9C3E06/2020\|EPI_ISL_566958\|20 | ···················· | ···················· | ···················· | ···················· | ···················· | ···················· | |
| K | 1980 | hCoV-19/England/MILK-9C397E/2020\|EPI_ISL_566959\|20 | ···················· | ···················· | ···················· | ···················· | ···················· | ···················· | |
| K | 1981 | hCoV-19/England/MILK-9C3B4B/2020\|EPI_ISL_566960\|20 | ···················· | ···················· | ···················· | ···················· | ···················· | ···················· | |
| K | 1982 | hCoV-19/England/MILK-9C3E9D/2020\|EPI_ISL_566961\|20 | ···················· | ·A·················· | ···················· | ···················· | ···················· | ···A················ | |
| K | 1983 | hCoV-19/England/MILK-9C3A8A/2020\|EPI_ISL_566962\|20 | ···················· | ·A·················· | ···················· | ···················· | ···················· | ···A················ | |
| K | 1984 | hCoV-19/England/MILK-9C3E33/2020\|EPI_ISL_566963\|20 | ···················· | ···················· | ···················· | ···················· | ···················· | ···················· | |
| K | 1985 | hCoV-19/England/MILK-9C338F/2020\|EPI_ISL_566965\|20 | ···················· | ···················· | ···················· | ···················· | ···················· | ···················· | |
| K | 1986 | hCoV-19/England/MILK-9C46D0/2020\|EPI_ISL_566966\|20 | ···················· | ···················· | ···················· | ···················· | ······T············· | ···················· | |
| K | 1987 | hCoV-19/England/MILK-9C45C4/2020\|EPI_ISL_566967\|20 | ···················· | ···················· | ···················· | ···················· | ···················· | ···················· | |
| K | 1988 | hCoV-19/England/MILK-9C4263/2020\|EPI_ISL_566968\|20 | ···················· | ···················· | ···················· | ···················· | ···················· | ···················· | |
| K | 1989 | hCoV-19/England/MILK-9C40F0/2020\|EPI_ISL_566969\|20 | ···················· | ···················· | ···················· | ···················· | ···················· | ···················· | |
| K | 1990 | hCoV-19/England/MILK-9C38EA/2020\|EPI_ISL_566971\|20 | ···················· | ···················· | ···················· | ···················· | ···················· | ···················· | |
| K | 1991 | hCoV-19/England/MILK-9C3844/2020\|EPI_ISL_566972\|20 | ···················· | ·A·················· | ···················· | ···················· | ···················· | ···A················ | |
| K | 1992 | hCoV-19/Wales/MILK-9C42AF/2020\|EPI_ISL_566974\|2020 | ···················· | ···················· | ···················· | ···················· | ···················· | ···················· | |
| K | 1993 | hCoV-19/England/MILK-9C311C/2020\|EPI_ISL_566975\|20 | ···················· | ···················· | ···················· | ···················· | ···················· | ···················· | |
| K | 1994 | hCoV-19/England/MILK-9C4649/2020\|EPI_ISL_566976\|20 | ···················· | ···················· | ···················· | ···················· | ···················· | ···················· | |
| K | 1995 | hCoV-19/England/MILK-9C41B1/2020\|EPI_ISL_566977\|20 | ···················· | ···················· | ···················· | ···················· | ···················· | ···················· | |
| K | 1996 | hCoV-19/England/MILK-9C3CDF/2020\|EPI_ISL_566978\|20 | ···················· | ···················· | ···················· | ···················· | ···················· | ···················· | |
| K | 1997 | hCoV-19/England/MILK-9C444F/2020\|EPI_ISL_566979\|20 | ···················· | ···················· | ···················· | ···················· | ···················· | ···················· | |
| K | 1998 | hCoV-19/England/MILK-9C33CB/2020\|EPI_ISL_566980\|20 | ···················· | ···················· | ···················· | ···················· | ···················· | ···················· | |
| K | 1999 | hCoV-19/Wales/MILK-9C44D6/2020\|EPI_ISL_566981\|2020 | ···················· | ···················· | ···················· | ···················· | ···················· | ···················· | |
| K | 2000 | hCoV-19/England/MILK-9C456A/2020\|EPI_ISL_566982\|20 | ···················· | ···················· | ···················· | ···················· | ···················· | ···················· | |
| K | 2001 | hCoV-19/England/MILK-9C400F/2020\|EPI_ISL_566983\|20 | ···················· | ···················· | ···················· | ···················· | ···················· | ···················· | |
| K | 2002 | hCoV-19/England/MILK-9C43F7/2020\|EPI_ISL_566984\|20 | ···················· | ···················· | ···················· | ···················· | ···················· | ···················· | |
| K | 2003 | hCoV-19/England/MILK-9C3A20/2020\|EPI_ISL_566985\|20 | ···················· | ······C············· | ···················· | ···················· | ···················· | ···················· | |
| K | 2004 | hCoV-19/France/PAC-IHU-1266/2020\|EPI_ISL_569100\|20 | ···················· | ···················· | ···················· | ···················· | ···················· | ···················· | |
| K | 2005 | hCoV-19/France/PAC-IHU-1267/2020\|EPI_ISL_569101\|20 | ···················· | ···················· | ···················· | ···················· | ···················· | ···················· | |
| K | 2006 | hCoV-19/France/PAC-IHU-1268/2020\|EPI_ISL_569102\|20 | ···················· | ···················· | ···················· | ···················· | ···················· | ···················· | |
| K | 2007 | hCoV-19/France/PAC-IHU-1270/2020\|EPI_ISL_569103\|20 | ···················· | ···················· | ···················· | ···················· | ···················· | ···················· | |
| K | 2008 | hCoV-19/France/PAC-IHU-1275/2020\|EPI_ISL_569104\|20 | ···················· | ···················· | ···················· | ···················· | ···················· | ···················· | |
| K | 2009 | hCoV-19/France/PAC-IHU-1279/2020\|EPI_ISL_569106\|20 | ···················· | ···················· | ···················· | ···················· | ···················· | ···················· | |
| K | 2010 | hCoV-19/France/PAC-IHU-1333/2020\|EPI_ISL_569120\|20 | ···················· | ···················· | ···················· | ···················· | ···················· | ···············A···· | |
| K | 2011 | hCoV-19/France/PAC-IHU-1334/2020\|EPI_ISL_569121\|20 | ···················· | ···················· | ···················· | ···················· | ···················· | ···················· | |
| K | 2012 | hCoV-19/France/PAC-IHU-1336/2020\|EPI_ISL_569123\|20 | ···················· | ···················· | ···················· | ···················· | ···················· | ···················· | |
| K | 2013 | hCoV-19/France/PAC-IHU-1337/2020\|EPI_ISL_569124\|20 | ···················· | ···················· | ···················· | ···················· | ···················· | ···················· | |
| K | 2014 | hCoV-19/France/PAC-IHU-1340/2020\|EPI_ISL_569125\|20 | ···················· | ···················· | ···················· | ···················· | ···················· | ···················· | |
| K | 2015 | hCoV-19/France/PAC-IHU-1341/2020\|EPI_ISL_569126\|20 | ···················· | ···················· | ···················· | ···················· | ···················· | ···················· | |
| K | 2016 | hCoV-19/France/PAC-IHU-1342/2020\|EPI_ISL_569127\|20 | ···················· | ···················· | ···················· | ···················· | ···················· | ···················· | |
| K | 2017 | hCoV-19/France/PAC-IHU-1344/2020\|EPI_ISL_569128\|20 | ···················· | ···················· | ···················· | ···················· | ···················· | ···················· | |

| | | | | | | | | |
|---|---|---|---|---|---|---|---|---|
| K | 2018 hCoV-19/France/PAC-IHU-1345/2020\|EPI_ISL_569129\|20 | ···················· | ···················· | ···················· | ···················· | ···················· | ···················· | |
| K | 2019 hCoV-19/France/PAC-IHU-1346/2020\|EPI_ISL_569130\|20 | ···················· | ···················· | ···················· | ···················· | ···················· | ···················· | |
| K | 2020 hCoV-19/France/PAC-IHU-1348/2020\|EPI_ISL_569132\|20 | ···················· | ···················· | ···················· | ···················· | ···················· | ···················· | |
| K | 2021 hCoV-19/France/PAC-IHU-1349/2020\|EPI_ISL_569133\|20 | ···················· | ···················· | ···················· | ···················· | ···················· | ···················· | |
| K | 2022 hCoV-19/France/PAC-IHU-1350/2020\|EPI_ISL_569134\|20 | ···················· | ···················· | ···················· | ···················· | ···················· | ···················· | |
| K | 2023 hCoV-19/France/PAC-IHU-1351/2020\|EPI_ISL_569135\|20 | ···················· | ··N················· | ···················· | ···················· | ···················· | ···················· | |
| K | 2024 hCoV-19/France/PAC-IHU-1353/2020\|EPI_ISL_569136\|20 | ···················· | ···················· | ···················· | ···················· | ···················· | ···················· | |
| K | 2025 hCoV-19/France/PAC-IHU-1355/2020\|EPI_ISL_569137\|20 | ···················· | ···················· | ···················· | ···················· | ···················· | ···················· | |
| K | 2026 hCoV-19/France/PAC-IHU-1357/2020\|EPI_ISL_569138\|20 | ···················· | ···················· | ···················· | ···················· | ···················· | ···················· | |
| K | 2027 hCoV-19/France/PAC-IHU-1359/2020\|EPI_ISL_569139\|20 | ···················· | ···················· | ···················· | ···················· | ···················· | ···················· | |
| K | 2028 hCoV-19/USA/ID-UW-6350/2020\|EPI_ISL_570100\|2020-04 | ···················· | ···················· | ···················· | ···················· | ···················· | ···················· | |
| K | 2029 hCoV-19/USA/WA-UW-6351/2020\|EPI_ISL_570101\|2020-04 | ···················· | ···················· | ···················· | ···················· | ···················· | ···················· | |
| K | 2030 hCoV-19/USA/WA-UW-6353/2020\|EPI_ISL_570102\|2020-04 | ···················· | ···················· | ···················· | ···················· | ···················· | ···················· | |
| K | 2031 hCoV-19/USA/WA-UW-6356/2020\|EPI_ISL_570103\|2020-04 | ···················· | ···················· | ···················· | ···················· | ···················· | ···················· | |
| K | 2032 hCoV-19/USA/CT-UW-6361/2020\|EPI_ISL_570104\|2020-04 | ···················· | ···················· | ···················· | ···················· | ···················· | ··DT···C·A········ | 1 |
| K | 2033 hCoV-19/USA/ID-UW-6365/2020\|EPI_ISL_570105\|2020-04 | ···················· | ···················· | ···················· | ···················· | ···················· | ···················· | |
| K | 2034 hCoV-19/USA/ID-UW-6367/2020\|EPI_ISL_570106\|2020-04 | ···················· | ···················· | ···················· | ···················· | ···················· | ···················· | |
| K | 2035 hCoV-19/USA/CT-UW-6369/2020\|EPI_ISL_570107\|2020-04 | ···················· | ···················· | ···················· | ···················· | ···················· | ···················· | |
| K | 2036 hCoV-19/USA/ID-UW-6377/2020\|EPI_ISL_570108\|2020-04 | ···················· | ···················· | ···················· | ···················· | ···················· | ···················· | |
| K | 2037 hCoV-19/USA/CT-UW-6379/2020\|EPI_ISL_570109\|2020-04 | ···················· | ···················· | ···················· | ···················· | ···················· | ···················· | |
| K | 2038 hCoV-19/USA/ID-UW-6383/2020\|EPI_ISL_570110\|2020-04 | ···················· | ···················· | ···················· | ···················· | ···················· | ···················· | |
| K | 2039 hCoV-19/USA/WA-UW-6406/2020\|EPI_ISL_570111\|2020-04 | ···················· | ···················· | ···················· | ···················· | ···················· | ···················· | |
| K | 2040 hCoV-19/USA/WA-UW-6413/2020\|EPI_ISL_570112\|2020-04 | ···················· | ···················· | ···················· | ···················· | ···················· | ···················· | |
| K | 2041 hCoV-19/USA/WA-UW-6438/2020\|EPI_ISL_570113\|2020-04 | ···················· | ···················· | ···················· | ···················· | ···················· | ···················· | |
| K | 2042 hCoV-19/USA/WA-UW-6439/2020\|EPI_ISL_570114\|2020-04 | ···················· | ···················· | ···················· | ···················· | ···················· | ···················· | |
| K | 2043 hCoV-19/USA/WA-UW-6441/2020\|EPI_ISL_570115\|2020-04 | ···················· | ···················· | ··T················· | ···················· | ···················· | ···················· | |
| K | 2044 hCoV-19/USA/WA-UW-6442/2020\|EPI_ISL_570116\|2020-04 | ···················· | ···················· | ···················· | ···················· | ···················· | ···················· | |
| K | 2045 hCoV-19/USA/ID-UW-6454/2020\|EPI_ISL_570117\|2020-04 | ···················· | ···················· | ···················· | ···················· | ···················· | ···················· | |
| K | 2046 hCoV-19/USA/ID-UW-6461/2020\|EPI_ISL_570118\|2020-04 | ···················· | ···················· | ···················· | ···················· | ···················· | ···················· | |
| K | 2047 hCoV-19/USA/ID-UW-6464/2020\|EPI_ISL_570119\|2020-04 | ···················· | ···················· | ···················· | ···················· | ···················· | ···················· | |
| K | 2048 hCoV-19/USA/ID-UW-6468/2020\|EPI_ISL_570120\|2020-04 | ···················· | ···················· | ···················· | ···················· | ···················· | ···················· | |
| K | 2049 hCoV-19/Australia/VIC10627/2020\|EPI_ISL_563000\|202 | ···················· | ···················· | ···················· | ···················· | ···················· | ···················· | |
| K | 2050 hCoV-19/Australia/VIC10631/2020\|EPI_ISL_563001\|202 | ···················· | ···················· | ···················· | ···················· | ···················· | ···················· | |
| K | 2051 hCoV-19/Australia/VIC10633/2020\|EPI_ISL_563002\|202 | ···················· | ···················· | ···················· | ···················· | ···················· | ···················· | |
| K | 2052 hCoV-19/Australia/VIC10636/2020\|EPI_ISL_563003\|202 | ···················· | ···················· | ···················· | ···················· | ···················· | ···················· | |
| K | 2053 hCoV-19/Australia/VIC10637/2020\|EPI_ISL_563004\|202 | ···················· | ···················· | ···················· | ···················· | ···················· | ···················· | |
| K | 2054 hCoV-19/Australia/VIC10638/2020\|EPI_ISL_563005\|202 | ···················· | ···················· | ···················· | ···················· | ···················· | ···················· | |
| K | 2056 hCoV-19/Australia/VIC10640/2020\|EPI_ISL_563007\|202 | ···················· | ···················· | ···················· | ···················· | ···················· | ···················· | |
| K | 2057 hCoV-19/Australia/VIC10643/2020\|EPI_ISL_563010\|202 | ···················· | ···················· | ···················· | ···················· | ···················· | ···················· | |
| K | 2058 hCoV-19/Australia/VIC10644/2020\|EPI_ISL_563011\|202 | ···················· | ···················· | ···················· | ···················· | ···················· | ···················· | |
| K | 2059 hCoV-19/Australia/VIC10645/2020\|EPI_ISL_563012\|202 | ···················· | ···················· | ···················· | ···················· | ···················· | ···················· | |
| K | 2060 hCoV-19/Australia/VIC10647/2020\|EPI_ISL_563013\|202 | ···················· | ···················· | ···················· | ···················· | ···················· | ···················· | |
| K | 2061 hCoV-19/Australia/VIC10648/2020\|EPI_ISL_563014\|202 | ···················· | ···················· | ···················· | ···················· | ···················· | ···················· | |
| K | 2062 hCoV-19/Australia/VIC10650/2020\|EPI_ISL_563015\|202 | ···················· | ···················· | ···················· | ···················· | ···················· | ···················· | |
| K | 2063 hCoV-19/Australia/VIC10651/2020\|EPI_ISL_563016\|202 | ···················· | ···················· | ···················· | ···················· | ···················· | ···················· | |
| K | 2064 hCoV-19/Australia/VIC10654/2020\|EPI_ISL_563017\|202 | ···················· | ···················· | ···················· | ···················· | ···················· | ···················· | |
| K | 2065 hCoV-19/Australia/VIC10655/2020\|EPI_ISL_563018\|202 | ···················· | ···················· | ···················· | ···················· | ···················· | ···················· | |
| K | 2066 hCoV-19/Australia/VIC10658/2020\|EPI_ISL_563019\|202 | ···················· | ···················· | ···················· | ···················· | ···················· | ···················· | |
| K | 2067 hCoV-19/Australia/VIC10659/2020\|EPI_ISL_563020\|202 | ···················· | ···················· | ···················· | ···················· | ···················· | ···················· | |
| K | 2068 hCoV-19/Australia/VIC10660/2020\|EPI_ISL_563021\|202 | ···················· | ···················· | ···················· | ···················· | ···················· | ···················· | |
| K | 2069 hCoV-19/Australia/VIC10661/2020\|EPI_ISL_563022\|202 | ···················· | ···················· | ···················· | ···················· | ···················· | ···················· | |
| K | 2070 hCoV-19/Australia/VIC10662/2020\|EPI_ISL_563023\|202 | ···················· | ···················· | ···················· | ···················· | ···················· | ···················· | |
| K | 2071 hCoV-19/Australia/VIC10663/2020\|EPI_ISL_563024\|202 | ···················· | ···················· | ···················· | ···················· | ···················· | ···················· | |
| K | 2073 hCoV-19/Australia/VIC10668/2020\|EPI_ISL_563026\|202 | ···················· | ···················· | ···················· | ···················· | ···················· | ···················· | |
| K | 2074 hCoV-19/Australia/VIC10671/2020\|EPI_ISL_563027\|202 | ···················· | ···················· | ···················· | ···················· | ···················· | ···················· | |
| K | 2075 hCoV-19/Australia/VIC10676/2020\|EPI_ISL_563028\|202 | ···················· | ···················· | ···················· | ···················· | ···················· | ···················· | |
| K | 2076 hCoV-19/Australia/VIC10680/2020\|EPI_ISL_563029\|202 | ···················· | ···················· | ···················· | ···················· | ···················· | ···················· | |
| K | 2077 hCoV-19/Australia/VIC10681/2020\|EPI_ISL_563030\|202 | ···················· | ···················· | ···················· | ···················· | ···················· | ···················· | |
| K | 2078 hCoV-19/Australia/VIC10682/2020\|EPI_ISL_563031\|202 | ···················· | ···················· | ···················· | ···················· | ···················· | ···················· | |
| K | 2079 hCoV-19/Australia/VIC10689/2020\|EPI_ISL_563032\|202 | ···················· | ···················· | ···················· | ···················· | ···················· | ···················· | |
| K | 2080 hCoV-19/Australia/VIC10690/2020\|EPI_ISL_563033\|202 | ···················· | ···················· | ···················· | ···················· | ···················· | ···················· | |
| K | 2081 hCoV-19/Australia/VIC10691/2020\|EPI_ISL_563034\|202 | ···················· | ···················· | ···················· | ···················· | ···················· | ···················· | |
| K | 2082 hCoV-19/Australia/VIC10694/2020\|EPI_ISL_563035\|202 | ···················· | ···················· | ···················· | ···················· | ···················· | ···················· | |
| K | 2083 hCoV-19/Australia/VIC10695/2020\|EPI_ISL_563036\|202 | ···················· | ···················· | ···················· | ···················· | ···················· | ···················· | |
| K | 2084 hCoV-19/Australia/VIC7272/2020\|EPI_ISL_564324\|2020 | ···················· | ···················· | ···················· | ···················· | ···················· | ···················· | |
| K | 2085 hCoV-19/Australia/VIC7275/2020\|EPI_ISL_564325\|2020 | ···················· | ···················· | ···················· | ···················· | ···················· | ···················· | |
| K | 2086 hCoV-19/Australia/VIC7276/2020\|EPI_ISL_564326\|2020 | ···················· | ···················· | ··C················· | ···················· | ···················· | ···················· | |
| K | 2087 hCoV-19/Australia/VIC7277/2020\|EPI_ISL_564327\|2020 | ···················· | ···················· | ···················· | ···················· | ···················· | ···················· | |
| K | 2088 hCoV-19/Australia/VIC7284/2020\|EPI_ISL_564328\|2020 | ···················· | ···················· | ···················· | ···················· | ···················· | ···················· | |
| K | 2089 hCoV-19/Australia/VIC7286/2020\|EPI_ISL_564329\|2020 | ···················· | ···················· | ···················· | ···················· | ···················· | ···················· | |
| K | 2090 hCoV-19/Australia/VIC7293/2020\|EPI_ISL_564330\|2020 | ···················· | ···················· | ···················· | ···················· | ···················· | ···················· | |
| K | 2091 hCoV-19/Australia/VIC7294/2020\|EPI_ISL_564331\|2020 | ···················· | ···················· | ···················· | ···················· | ···················· | ···················· | |
| K | 2092 hCoV-19/Australia/VIC7299/2020\|EPI_ISL_564332\|2020 | ···················· | ···················· | ···················· | ···················· | ···················· | ···················· | |
| K | 2093 hCoV-19/Australia/VIC7302/2020\|EPI_ISL_564333\|2020 | ···················· | ···················· | ···················· | ···················· | ···················· | ···················· | |
| K | 2094 hCoV-19/Australia/VIC7303/2020\|EPI_ISL_564334\|2020 | ···················· | ···················· | ···················· | ···················· | ···················· | ···················· | |
| K | 2095 hCoV-19/Australia/VIC7305/2020\|EPI_ISL_564335\|2020 | ···················· | ···················· | ···················· | ···················· | ···················· | ···················· | |
| K | 2096 hCoV-19/Australia/VIC7306/2020\|EPI_ISL_564336\|2020 | ···················· | ···················· | ···················· | ···················· | ···················· | ···················· | |
| K | 2097 hCoV-19/Australia/VIC7308/2020\|EPI_ISL_564337\|2020 | ···················· | ···················· | ···················· | ···················· | ···················· | ···················· | |
| K | 2098 hCoV-19/Australia/VIC7309/2020\|EPI_ISL_564338\|2020 | ···················· | ···················· | ···················· | ···················· | ···················· | ···················· | |
| K | 2099 hCoV-19/Australia/VIC7316/2020\|EPI_ISL_564340\|2020 | ···················· | ·····A·············· | ···················· | ···················· | ···················· | ···················· | |

| | | | | | | | | |
|---|---|---|---|---|---|---|---|---|
| K | 2100 hCoV-19/Australia/VIC7317/2020\|EPI_ISL_564341\|2020 | ···················· | ···················· | ···················· | ···················· | ···················· | ···················· | |
| K | 2101 hCoV-19/Australia/VIC7323/2020\|EPI_ISL_564342\|2020 | ···················· | ···················· | ···················· | ···················· | ···················· | ···················· | |
| K | 2102 hCoV-19/Australia/VIC7325/2020\|EPI_ISL_564343\|2020 | ···················· | ···················· | ···················· | ···················· | ···················· | ···················· | |
| K | 2103 hCoV-19/Australia/VIC7327/2020\|EPI_ISL_564344\|2020 | ···················· | ···················· | ···················· | ···················· | ···················· | ···················· | |
| K | 2104 hCoV-19/Australia/VIC7328/2020\|EPI_ISL_564345\|2020 | ···················· | ···················· | ···················· | ···················· | ···················· | ···················· | |
| K | 2105 hCoV-19/Australia/VIC7334/2020\|EPI_ISL_564346\|2020 | ···················· | ···················· | ···················· | ···················· | ···················· | ···················· | |
| K | 2106 hCoV-19/Australia/VIC7337/2020\|EPI_ISL_564347\|2020 | ···················· | ···················· | ···················· | ···················· | ···················· | ···················· | |
| K | 2107 hCoV-19/Australia/VIC7339/2020\|EPI_ISL_564348\|2020 | ···················· | ···················· | ···················· | ···················· | ···················· | ···················· | |
| K | 2108 hCoV-19/Australia/VIC7343/2020\|EPI_ISL_564349\|2020 | ···················· | ···················· | ···················· | ···················· | ···················· | ···················· | |
| K | 2109 hCoV-19/Australia/VIC7344/2020\|EPI_ISL_564350\|2020 | ···················· | ···················· | ···················· | ···················· | ···················· | ···················· | |
| K | 2110 hCoV-19/Australia/VIC7345/2020\|EPI_ISL_564351\|2020 | ···················· | ···················· | ···················· | ···················· | ···················· | ···················· | |
| K | 2111 hCoV-19/Australia/VIC7348/2020\|EPI_ISL_564352\|2020 | ···················· | ···················· | ···K················ | ···················· | ···················· | ···················· | |
| K | 2112 hCoV-19/Australia/VIC7349/2020\|EPI_ISL_564353\|2020 | ···················· | ···················· | ···················· | ···················· | ···················· | ···················· | |
| K | 2113 hCoV-19/Australia/VIC7350/2020\|EPI_ISL_564354\|2020 | ···················· | ···················· | ···················· | ···················· | ···················· | ···················· | |
| K | 2114 hCoV-19/Australia/VIC7351/2020\|EPI_ISL_564355\|2020 | ···················· | ···················· | ···················· | ···················· | ···················· | ···················· | |
| K | 2115 hCoV-19/Australia/VIC7352/2020\|EPI_ISL_564356\|2020 | ···················· | ···················· | ···················· | ···················· | ···················· | ···················· | |
| K | 2116 hCoV-19/Australia/VIC7355/2020\|EPI_ISL_564357\|2020 | ···················· | ···················· | ···K················ | ···················· | ···················· | ···················· | |
| K | 2117 hCoV-19/Australia/VIC7360/2020\|EPI_ISL_564358\|2020 | ···················· | ···················· | ···················· | ···················· | ···················· | ···················· | |
| K | 2118 hCoV-19/Australia/VIC7362/2020\|EPI_ISL_564359\|2020 | ···················· | ···················· | ···················· | ···················· | ···················· | ···················· | |
| K | 2119 hCoV-19/Australia/VIC7363/2020\|EPI_ISL_564360\|2020 | ···················· | ···················· | ···················· | ···················· | ···················· | ···················· | |
| K | 2120 hCoV-19/Australia/VIC7364/2020\|EPI_ISL_564361\|2020 | ···················· | ···················· | ···················· | ···················· | ···················· | ···················· | |
| K | 2121 hCoV-19/Australia/VIC7365/2020\|EPI_ISL_564362\|2020 | ···················· | ···················· | ···················· | ···················· | ···················· | ···················· | |
| K | 2122 hCoV-19/Australia/VIC7367/2020\|EPI_ISL_564363\|2020 | ···················· | ···················· | ···················· | ···················· | ···················· | ···················· | |
| K | 2123 hCoV-19/Australia/VIC7368/2020\|EPI_ISL_564364\|2020 | ···················· | ···················· | ···················· | ···················· | ···················· | ···················· | |
| K | 2124 hCoV-19/Australia/VIC7372/2020\|EPI_ISL_564365\|2020 | ···················· | ···················· | ···················· | ···················· | ···················· | ···················· | |
| K | 2125 hCoV-19/Australia/VIC7373/2020\|EPI_ISL_564366\|2020 | ···················· | ···················· | ···················· | ···················· | ···················· | ···················· | |
| K | 2126 hCoV-19/Australia/VIC7374/2020\|EPI_ISL_564367\|2020 | ···················· | ···················· | ···················· | ···················· | ···················· | ···················· | |
| K | 2127 hCoV-19/Australia/VIC9689/2020\|EPI_ISL_565655\|2020 | ···················· | ···················· | ···················· | ···················· | ···················· | ···················· | |
| K | 2128 hCoV-19/Australia/VIC9691/2020\|EPI_ISL_565656\|2020 | ···················· | ···················· | ···················· | ···················· | ···················· | ···················· | |
| K | 2129 hCoV-19/Australia/VIC9694/2020\|EPI_ISL_565657\|2020 | ···················· | ···················· | ···················· | ···················· | ···················· | ···················· | |
| K | 2130 hCoV-19/Australia/VIC9695/2020\|EPI_ISL_565658\|2020 | ···················· | ···················· | ···················· | ···················· | ···················· | ···················· | |
| K | 2131 hCoV-19/Australia/VIC9696/2020\|EPI_ISL_565659\|2020 | ···················· | ···················· | ···················· | ···················· | ···················· | ···················· | |
| K | 2132 hCoV-19/Australia/VIC9697/2020\|EPI_ISL_565660\|2020 | ···················· | ···················· | ···················· | ···················· | ···················· | ···················· | |
| K | 2133 hCoV-19/Australia/VIC9698/2020\|EPI_ISL_565661\|2020 | ···················· | ···················· | ···················· | ···················· | ···················· | ···················· | |
| K | 2134 hCoV-19/Australia/VIC9700/2020\|EPI_ISL_565662\|2020 | ···················· | ···················· | ···················· | ···················· | ···················· | ···················· | |
| K | 2135 hCoV-19/Australia/VIC9705/2020\|EPI_ISL_565663\|2020 | ···················· | ···················· | ···················· | ···················· | ···················· | ···················· | |
| K | 2136 hCoV-19/Australia/VIC9707/2020\|EPI_ISL_565664\|2020 | ···················· | ···················· | ···················· | ···················· | ···················· | ···················· | |
| K | 2137 hCoV-19/Australia/VIC9708/2020\|EPI_ISL_565665\|2020 | ···················· | ···················· | ···················· | ···················· | ···················· | ···················· | |
| K | 2138 hCoV-19/Australia/VIC9711/2020\|EPI_ISL_565666\|2020 | ···················· | ···················· | ···················· | ···················· | ···················· | ···················· | |
| K | 2139 hCoV-19/Australia/VIC9713/2020\|EPI_ISL_565667\|2020 | ···················· | ···················· | ···················· | ···················· | ···················· | ···················· | |
| K | 2142 hCoV-19/Australia/VIC9716/2020\|EPI_ISL_565670\|2020 | ···················· | ···················· | ···················· | ···················· | ···················· | ···················· | |
| K | 2143 hCoV-19/Australia/VIC9717/2020\|EPI_ISL_565671\|2020 | ···················· | ···················· | ···················· | ···················· | ···················· | ···················· | |
| K | 2145 hCoV-19/Australia/VIC9722/2020\|EPI_ISL_565673\|2020 | ···················· | ···················· | ···················· | ···················· | ···················· | ···················· | |
| K | 2146 hCoV-19/Australia/VIC9724/2020\|EPI_ISL_565674\|2020 | ···················· | ···················· | ···················· | ···················· | ···················· | ···················· | |
| K | 2147 hCoV-19/Australia/VIC9725/2020\|EPI_ISL_565675\|2020 | ···················· | ···················· | ···················· | ···················· | ···················· | ···················· | |
| K | 2148 hCoV-19/Australia/VIC9726/2020\|EPI_ISL_565676\|2020 | ···················· | ···················· | ···················· | ···················· | ···················· | ···················· | |
| K | 2149 hCoV-19/Australia/VIC9728/2020\|EPI_ISL_565677\|2020 | ···················· | ···················· | ···················· | ···················· | ···················· | ···················· | |
| K | 2150 hCoV-19/Australia/VIC9729/2020\|EPI_ISL_565678\|2020 | ···················· | ···················· | ···················· | ···················· | ···················· | ···················· | |
| K | 2151 hCoV-19/Australia/VIC9730/2020\|EPI_ISL_565679\|2020 | ···················· | ···················· | ···················· | ···················· | ···················· | ···················· | |
| K | 2154 hCoV-19/Australia/VIC9735/2020\|EPI_ISL_565682\|2020 | ···················· | ···················· | ···················· | ···················· | ···················· | ···················· | |
| K | 2156 hCoV-19/Australia/VIC9738/2020\|EPI_ISL_565684\|2020 | ···················· | ···················· | ···················· | ···················· | ···················· | ···················· | |
| K | 2157 hCoV-19/Australia/VIC9739/2020\|EPI_ISL_565685\|2020 | ···················· | ···················· | ···················· | ···················· | ···················· | ···················· | |
| K | 2161 hCoV-19/Australia/VIC9746/2020\|EPI_ISL_565689\|2020 | ···················· | ···················· | ···················· | ···················· | NNN················· | ···················· | 1 |
| K | 2162 hCoV-19/Australia/VIC9748/2020\|EPI_ISL_565690\|2020 | ···················· | ···················· | ···················· | ···················· | ···················· | ···················· | |
| K | 2163 hCoV-19/Australia/VIC9749/2020\|EPI_ISL_565691\|2020 | ···················· | ···················· | ···················· | ···················· | ···················· | ···················· | |
| K | 2164 hCoV-19/Australia/VIC9750/2020\|EPI_ISL_565692\|2020 | ···················· | ···················· | ···················· | ···················· | ···················· | ···················· | |
| K | 2165 hCoV-19/Australia/VIC9752/2020\|EPI_ISL_565693\|2020 | ···················· | ···················· | ···················· | ···················· | ···················· | ···················· | |
| K | 2166 hCoV-19/Australia/VIC9756/2020\|EPI_ISL_565694\|2020 | ···················· | ···················· | ···················· | ···················· | ···················· | ···················· | |
| K | 2167 hCoV-19/Australia/VIC9758/2020\|EPI_ISL_565695\|2020 | ···················· | ···················· | ···················· | ···················· | ···················· | ···················· | |
| K | 2168 hCoV-19/Australia/VIC9760/2020\|EPI_ISL_565696\|2020 | ···················· | ···················· | ···················· | ···················· | N··················· | ···················· | |
| K | 2169 hCoV-19/Australia/VIC9761/2020\|EPI_ISL_565697\|2020 | ···················· | ···················· | ···················· | ···················· | ···················· | ···················· | |
| K | 2171 hCoV-19/England/MILK-9C447C/2020\|EPI_ISL_566900\|20 | ···················· | ···················· | ···················· | ···················· | ···················· | ···················· | |
| K | 2172 hCoV-19/England/MILK-9C4184/2020\|EPI_ISL_566901\|20 | ···················· | ···················· | ···················· | ···················· | ···················· | ···················· | |
| K | 2173 hCoV-19/Wales/MILK-9C3158/2020\|EPI_ISL_566902\|2020 | ···················· | ···················· | ···················· | ···················· | ···················· | ···················· | |
| K | 2174 hCoV-19/England/MILK-9C39C9/2020\|EPI_ISL_566903\|20 | ···················· | ···················· | ···················· | ···················· | ···················· | ···················· | |
| K | 2175 hCoV-19/England/MILK-9C2FF5/2020\|EPI_ISL_566906\|20 | ···················· | ···················· | ···················· | ···················· | ···················· | ···················· | |
| K | 2176 hCoV-19/England/MILK-9C306A/2020\|EPI_ISL_566907\|20 | ···················· | ···················· | ···················· | ···················· | ···················· | ···················· | |
| K | 2177 hCoV-19/England/MILK-9C3E7F/2020\|EPI_ISL_566908\|20 | ···················· | ···················· | ···················· | ···················· | ···················· | ···················· | |
| K | 2178 hCoV-19/Wales/MILK-9C3D36/2020\|EPI_ISL_566909\|2020 | ···················· | ···················· | ···················· | ···················· | ···················· | ···················· | |
| K | 2179 hCoV-19/England/MILK-9C36FF/2020\|EPI_ISL_566910\|20 | ···················· | ···················· | ···················· | ···················· | ···················· | ···················· | |
| K | 2180 hCoV-19/England/MILK-9C31FE/2020\|EPI_ISL_566911\|20 | ···················· | ···················· | ···················· | ···················· | ···················· | ···················· | |
| K | 2181 hCoV-19/England/MILK-9C45F1/2020\|EPI_ISL_566912\|20 | ···················· | ···················· | ···················· | ···················· | ···················· | ···················· | |
| K | 2182 hCoV-19/England/MILK-9C3FD6/2020\|EPI_ISL_566913\|20 | ···················· | ···················· | ···················· | ···················· | ······T············· | ···················· | |
| K | 2183 hCoV-19/England/MILK-9C364A/2020\|EPI_ISL_566914\|20 | ···················· | ···················· | ···················· | ···················· | ···················· | ···················· | |
| K | 2184 hCoV-19/England/MILK-9C34F5/2020\|EPI_ISL_566915\|20 | ···················· | ···················· | ···················· | ···················· | ···················· | ···················· | |
| K | 2185 hCoV-19/Wales/MILK-9C37FC/2020\|EPI_ISL_566916\|2020 | ···················· | ···················· | ···················· | ···················· | ···················· | ···················· | |
| K | 2186 hCoV-19/England/MILK-9C3C2A/2020\|EPI_ISL_566917\|20 | ···················· | ···················· | ···················· | ···················· | ···················· | ···················· | |
| K | 2187 hCoV-19/England/MILK-9C4421/2020\|EPI_ISL_566918\|20 | ···················· | ···················· | ···················· | ···················· | ···················· | ···················· | |
| K | 2188 hCoV-19/England/MILK-9C304C/2020\|EPI_ISL_566919\|20 | ···················· | ···················· | ···················· | ···················· | ···················· | ···················· | |
| K | 2189 hCoV-19/England/MILK-9C45A6/2020\|EPI_ISL_566987\|20 | ···················· | ···················· | ···················· | ···················· | ···················· | ···················· | |

| | | | | | | | | |
|---|---|---|---|---|---|---|---|---|
| K | 2190 | hCoV-19/England/MILK-9C3F8B/2020\|EPI_ISL_566988\|20 | ···················· | ···················· | ···················· | ···················· | ···················· | ···················· |
| K | 2191 | hCoV-19/England/MILK-9C2F40/2020\|EPI_ISL_566989\|20 | ···················· | ···················· | ···················· | ···················· | ···················· | ···················· |
| K | 2192 | hCoV-19/England/MILK-9C4175/2020\|EPI_ISL_566991\|20 | ···················· | ···················· | ···················· | ···················· | ···················· | ···················· |
| K | 2193 | hCoV-19/Wales/MILK-9C352F/2020\|EPI_ISL_566992\|2020 | ···················· | ···················· | ···················· | ···················· | ···················· | ···················· |
| K | 2194 | hCoV-19/England/MILK-9C4430/2020\|EPI_ISL_566993\|20 | ···················· | ···················· | ···················· | ···················· | ···················· | ···················· |
| K | 2195 | hCoV-19/England/MILK-9C3316/2020\|EPI_ISL_566994\|20 | ···················· | ···················· | ···················· | ···················· | ···················· | ···················· |
| K | 2196 | hCoV-19/England/MILK-9C3DCD/2020\|EPI_ISL_566995\|20 | ···················· | ···················· | ···················· | ···················· | ···················· | ···················· |
| K | 2197 | hCoV-19/England/MILK-9C4236/2020\|EPI_ISL_566996\|20 | ···················· | ···················· | ···················· | ···················· | ···················· | ···················· |
| K | 2198 | hCoV-19/Wales/MILK-9C3DBE/2020\|EPI_ISL_566997\|2020 | ···················· | ···················· | ···················· | ···················· | ···················· | ···················· |
| K | 2199 | hCoV-19/England/QEUH-9C8180/2020\|EPI_ISL_566998\|20 | ···················· | ···················· | ···················· | ···················· | ···················· | ···················· |
| K | 2200 | hCoV-19/England/QEUH-9C850C/2020\|EPI_ISL_566999\|20 | ···················· | ···················· | ···················· | ···················· | ···················· | ···················· |
| K | 2201 | hCoV-19/France/PAC-IHU-1360/2020\|EPI_ISL_569140\|20 | ···················· | ···················· | ···················· | ···················· | ···················· | ···················· |
| K | 2202 | hCoV-19/France/PAC-IHU-1362/2020\|EPI_ISL_569141\|20 | ···················· | ···················· | ···················· | ···················· | ···················· | ···················· |
| K | 2203 | hCoV-19/France/PAC-IHU-1363/2020\|EPI_ISL_569142\|20 | ···················· | ···················· | ···················· | ···················· | ···················· | ···················· |
| K | 2204 | hCoV-19/France/PAC-IHU-1364/2020\|EPI_ISL_569143\|20 | ···················· | ···················· | ···················· | ···················· | ···················· | ···················· |
| K | 2205 | hCoV-19/France/PAC-IHU-1365/2020\|EPI_ISL_569144\|20 | ···················· | ···················· | ···················· | ···················· | ···················· | ···················· |
| K | 2206 | hCoV-19/France/PAC-IHU-1374/2020\|EPI_ISL_569145\|20 | ···················· | ·A·················· | ···················· | ···················· | ···················· | ···················· |
| K | 2207 | hCoV-19/France/PAC-IHU-1377/2020\|EPI_ISL_569146\|20 | ···················· | ···················· | ···················· | ···················· | ···················· | ···················· |
| K | 2208 | hCoV-19/France/PAC-IHU-1382/2020\|EPI_ISL_569147\|20 | ···················· | ····A··············· | ···················· | ···················· | ···················· | ···················· |
| K | 2209 | hCoV-19/France/PAC-IHU-1387/2020\|EPI_ISL_569149\|20 | ···················· | ···················· | ···················· | ···················· | ·····N·············· | ···················· |
| K | 2210 | hCoV-19/France/PAC-IHU-1388/2020\|EPI_ISL_569150\|20 | ···················· | ···················· | ···················· | ···················· | ···················· | ···················· |
| K | 2211 | hCoV-19/France/PAC-IHU-1390/2020\|EPI_ISL_569151\|20 | ···················· | ···················· | ···················· | ···················· | ···················· | ···················· |
| K | 2212 | hCoV-19/France/PAC-IHU-1410/2020\|EPI_ISL_569155\|20 | ···················· | ···················· | ···················· | ···················· | ···················· | ···················· |
| K | 2213 | hCoV-19/France/PAC-IHU-1414/2020\|EPI_ISL_569156\|20 | ···················· | ···················· | ···················· | ···················· | ···················· | ···················· |
| K | 2214 | hCoV-19/France/PAC-IHU-1419/2020\|EPI_ISL_569157\|20 | ···················· | ·A·················· | ···················· | ···················· | ···················· | ···················· |
| K | 2215 | hCoV-19/France/PAC-IHU-1420/2020\|EPI_ISL_569158\|20 | ···················· | ···················· | ···················· | ···················· | ···················· | ···················· |
| K | 2216 | hCoV-19/France/PAC-IHU-1421/2020\|EPI_ISL_569159\|20 | ···················· | ···················· | ···················· | ···················· | ···················· | ···················· |
| K | 2217 | hCoV-19/France/PAC-IHU-1426/2020\|EPI_ISL_569160\|20 | ···················· | ···················· | ···················· | ···················· | ···················· | ···················· |
| K | 2218 | hCoV-19/France/PAC-IHU-1427/2020\|EPI_ISL_569161\|20 | ···················· | ···················· | ···················· | ···················· | ···················· | ···················· |
| K | 2219 | hCoV-19/France/PAC-IHU-1428/2020\|EPI_ISL_569162\|20 | ···················· | ···················· | ···················· | ···················· | ···················· | ···················· |
| K | 2220 | hCoV-19/France/PAC-IHU-1430/2020\|EPI_ISL_569164\|20 | ···················· | ·A·················· | ···················· | ···················· | ···················· | ···················· |
| K | 2221 | hCoV-19/France/PAC-IHU-1431/2020\|EPI_ISL_569165\|20 | ···················· | ···················· | ···················· | ···················· | ···················· | ···················· |
| K | 2222 | hCoV-19/France/PAC-IHU-1432/2020\|EPI_ISL_569166\|20 | ···················· | ·A·················· | ···················· | ···················· | ···················· | ···················· |
| K | 2223 | hCoV-19/France/PAC-IHU-1438/2020\|EPI_ISL_569168\|20 | ···················· | ···················· | ···················· | ···················· | ···················· | ···················· |
| K | 2224 | hCoV-19/France/PAC-IHU-1439/2020\|EPI_ISL_569169\|20 | ···················· | ···················· | ···················· | ···················· | ···················· | ···················· |
| K | 2225 | hCoV-19/France/PAC-IHU-1446/2020\|EPI_ISL_569170\|20 | ···················· | ···················· | ···················· | ···················· | ···················· | ···················· |
| K | 2226 | hCoV-19/France/PAC-IHU-1448/2020\|EPI_ISL_569171\|20 | ···················· | ···················· | ···················· | ···················· | ···················· | ···················· |
| K | 2227 | hCoV-19/France/PAC-IHU-1461/2020\|EPI_ISL_569172\|20 | ···················· | ·A·················· | ···················· | ···················· | ···················· | ···················· |
| K | 2229 | hCoV-19/France/PAC-IHU-1470/2020\|EPI_ISL_569174\|20 | ···················· | ···················· | ···················· | ···················· | ···················· | ···················· |
| K | 2230 | hCoV-19/France/PAC-IHU-1475/2020\|EPI_ISL_569175\|20 | ···················· | ···················· | ···················· | ···················· | ···················· | ···················· |
| K | 2231 | hCoV-19/France/PAC-IHU-1476/2020\|EPI_ISL_569176\|20 | ···················· | ·A·················· | ···················· | ···················· | ···················· | ···················· |
| K | 2232 | hCoV-19/France/PAC-IHU-1481/2020\|EPI_ISL_569177\|20 | ···················· | ···················· | ···················· | ···················· | ···················· | ···················· |
| K | 2233 | hCoV-19/France/PAC-IHU-1486/2020\|EPI_ISL_569178\|20 | ···················· | ···················· | ···················· | ···················· | ···················· | ···················· |
| K | 2234 | hCoV-19/France/PAC-IHU-1488/2020\|EPI_ISL_569179\|20 | ···················· | ···················· | ···················· | ···················· | ···················· | ···················· |
| K | 2235 | hCoV-19/France/PAC-IHU-1502/2020\|EPI_ISL_569180\|20 | ···················· | ·A·················· | ···················· | ···················· | ···················· | ···················· |
| K | 2236 | hCoV-19/France/PAC-IHU-1523/2020\|EPI_ISL_569187\|20 | ···················· | ····A··············· | ···················· | ···················· | ···················· | ···················· |
| K | 2237 | hCoV-19/France/PAC-IHU-1524/2020\|EPI_ISL_569188\|20 | ···················· | ···················· | ···················· | ···················· | ···················· | ···················· |
| K | 2238 | hCoV-19/France/PAC-IHU-1525/2020\|EPI_ISL_569189\|20 | ···················· | ···················· | ···················· | ···················· | ···················· | ···················· |
| K | 2239 | hCoV-19/France/PAC-IHU-1527/2020\|EPI_ISL_569190\|20 | ···················· | ···················· | ···················· | ···················· | ···················· | ···················· |
| K | 2240 | hCoV-19/France/PAC-IHU-1529/2020\|EPI_ISL_569191\|20 | ···················· | ···················· | ···················· | ···················· | ···················· | ···················· |
| K | 2241 | hCoV-19/France/PAC-IHU-1540/2020\|EPI_ISL_569192\|20 | ········T··········· | ···················· | ···················· | ···················· | ···················· | ···················· |
| K | 2242 | hCoV-19/France/PAC-IHU-1548/2020\|EPI_ISL_569193\|20 | ···················· | ···················· | ···················· | ···················· | ···················· | ···················· |
| K | 2243 | hCoV-19/France/PAC-IHU-1552/2020\|EPI_ISL_569195\|20 | ···················· | ···················· | ···················· | ···················· | ···················· | ··N················· |
| K | 2244 | hCoV-19/France/PAC-IHU-1560/2020\|EPI_ISL_569196\|20 | ···················· | ···················· | ···················· | ···················· | ···················· | ···················· |
| K | 2245 | hCoV-19/France/PAC-IHU-1564/2020\|EPI_ISL_569197\|20 | ···················· | ···················· | ···················· | ···················· | ···················· | ···················· |
| K | 2246 | hCoV-19/France/PAC-IHU-1565/2020\|EPI_ISL_569198\|20 | ···················· | ···················· | ···················· | ···················· | ···················· | ···················· |
| K | 2247 | hCoV-19/France/PAC-IHU-1566/2020\|EPI_ISL_569199\|20 | ···················· | ···················· | ···················· | ···················· | ···················· | ···················· |
| K | 2248 | hCoV-19/USA/ID-UW-6469/2020\|EPI_ISL_570121\|2020-04 | ···················· | ···················· | ···················· | ···················· | ···················· | ···················· |
| K | 2249 | hCoV-19/USA/ID-UW-6470/2020\|EPI_ISL_570122\|2020-04 | ···················· | ···················· | ···················· | ···················· | ···················· | ···················· |
| K | 2250 | hCoV-19/USA/WA-UW-6496/2020\|EPI_ISL_570123\|2020-04 | ···················· | ···················· | ···················· | ···················· | ···················· | ···················· |
| K | 2251 | hCoV-19/USA/WA-UW-6502/2020\|EPI_ISL_570124\|2020-04 | ···················· | ···················· | ···················· | ···················· | ···················· | ···················· |
| K | 2252 | hCoV-19/USA/WA-UW-6511/2020\|EPI_ISL_570125\|2020-04 | ···················· | ···················· | ···················· | ···················· | ···················· | ···················· |
| K | 2253 | hCoV-19/USA/WA-UW-6525/2020\|EPI_ISL_570126\|2020-04 | ···················· | ···················· | ···················· | ···················· | ···················· | ···················· |
| K | 2254 | hCoV-19/USA/WA-UW-6543/2020\|EPI_ISL_570127\|2020-04 | ···················· | ···················· | ···················· | ···················· | ···················· | ···················· |
| K | 2255 | hCoV-19/USA/WA-UW-6550/2020\|EPI_ISL_570128\|2020-04 | ···················· | ···················· | ···················· | ···················· | ···················· | ···················· |
| K | 2256 | hCoV-19/USA/WA-UW-6560/2020\|EPI_ISL_570129\|2020-04 | ···················· | ···················· | ···················· | ···················· | ···················· | ···················· |
| K | 2257 | hCoV-19/USA/CT-UW-6563/2020\|EPI_ISL_570130\|2020-04 | ···················· | ···················· | ···················· | ···················· | ···················· | ···················· |
| K | 2258 | hCoV-19/USA/CT-UW-6566/2020\|EPI_ISL_570131\|2020-04 | ···················· | ···················· | ···················· | ···················· | ···················· | ···················· |
| K | 2259 | hCoV-19/USA/CT-UW-6572/2020\|EPI_ISL_570132\|2020-04 | ···················· | ····A··············· | ···················· | ···················· | ···················· | ···················· |
| K | 2260 | hCoV-19/USA/CT-UW-6684/2020\|EPI_ISL_570133\|2020-04 | ···················· | ···················· | ···················· | ···················· | ···················· | ···················· |
| K | 2261 | hCoV-19/USA/CT-UW-6691/2020\|EPI_ISL_570135\|2020-04 | ···················· | ···················· | ···················· | ···················· | ···················· | ···················· |
| K | 2262 | hCoV-19/USA/WA-UW-6706/2020\|EPI_ISL_570136\|2020-04 | ···················· | ···················· | ···················· | ···················· | ···················· | ···················· |
| K | 2263 | hCoV-19/USA/CT-UW-6709/2020\|EPI_ISL_570137\|2020-04 | ···················· | ···················· | ···················· | ···················· | ···················· | ···················· |
| K | 2264 | hCoV-19/USA/WA-UW-6716/2020\|EPI_ISL_570138\|2020-04 | ···················· | ···················· | ···················· | ···················· | ···················· | ···················· |
| K | 2265 | hCoV-19/USA/WA-UW-6718/2020\|EPI_ISL_570139\|2020-04 | ···················· | ···················· | ···················· | ···················· | ···················· | ···················· |
| K | 2266 | hCoV-19/USA/WA-UW-6750/2020\|EPI_ISL_570140\|2020-04 | ···················· | ···················· | ···················· | ···················· | ···················· | ···················· |
| K | 2267 | hCoV-19/USA/CT-UW-6758/2020\|EPI_ISL_570141\|2020-04 | ···················· | ···················· | ···················· | ···················· | ···················· | ···················· |
| K | 2268 | hCoV-19/USA/WA-UW-6762/2020\|EPI_ISL_570142\|2020-04 | ···················· | ···················· | ···················· | ···················· | ···················· | ···················· |
| K | 2269 | hCoV-19/USA/WA-UW-6765/2020\|EPI_ISL_570143\|2020-04 | ···················· | ···················· | ···················· | ···················· | ···················· | ···················· |
| K | 2270 | hCoV-19/USA/WA-UW-6767/2020\|EPI_ISL_570144\|2020-04 | ···················· | ···················· | ···················· | ···················· | ···················· | ···················· |

| | | | | | | | | | |
|---|---|---|---|---|---|---|---|---|---|
| K | 2271 | hCoV-19/USA/CT-UW-6780/2020\|EPI_ISL_570145\|2020-04 | ···················· | ···················A | ···················· | ···················· | ···················· | ···················· | 1 |
| K | 2272 | hCoV-19/USA/CT-UW-6790/2020\|EPI_ISL_570146\|2020-04 | ···················· | ···················· | ···················· | ···················· | ···················· | ···················· | |
| K | 2273 | hCoV-19/USA/WA-UW-6806/2020\|EPI_ISL_570147\|2020-04 | ···················· | ···················· | ···················· | ···················· | ···················· | ···················· | |
| K | 2274 | hCoV-19/USA/CT-UW-6807/2020\|EPI_ISL_570148\|2020-04 | ···················· | ···················· | ···················· | ···················· | ···················· | ···················· | |
| K | 2275 | hCoV-19/USA/WA-UW-6820/2020\|EPI_ISL_570150\|2020-04 | ···················· | ···················· | ···················· | ···················· | ···················· | ···················· | |
| K | 2276 | hCoV-19/USA/WA-UW-6822/2020\|EPI_ISL_570151\|2020-04 | ···················· | ···················· | ···················· | ···················· | ···················· | ···················· | |
| K | 2277 | hCoV-19/USA/WA-UW-6971/2020\|EPI_ISL_570152\|2020-03 | ···················· | ···················· | ···················· | ···················· | ···················· | ···················· | |
| K | 2278 | hCoV-19/USA/WA-UW-6976/2020\|EPI_ISL_570153\|2020-03 | ···················· | ···················· | ···················· | ···················· | ···················· | ···················· | |
| K | 2279 | hCoV-19/USA/WA-UW-6980/2020\|EPI_ISL_570154\|2020-03 | ···················· | ···················· | ···················· | ···················· | ···················· | ···················· | |
| K | 2280 | hCoV-19/USA/WA-UW-6985/2020\|EPI_ISL_570155\|2020-03 | ···················· | ···················· | ···················· | ···················· | ···················· | ···················· | |
| K | 2281 | hCoV-19/USA/WA-UW-6990/2020\|EPI_ISL_570156\|2020-03 | ···················· | ···················· | ···················· | ···················· | ···················· | ···················· | |
| K | 2282 | hCoV-19/USA/WA-UW-7006/2020\|EPI_ISL_570157\|2020-04 | ···················· | ···················· | ···················· | ···················· | ···················· | ···················· | |
| K | 2283 | hCoV-19/USA/WA-UW-7007/2020\|EPI_ISL_570158\|2020-04 | ···················· | ···················· | ···················· | ···················· | ···················· | ···················· | |
| K | 2284 | hCoV-19/USA/WA-UW-7011/2020\|EPI_ISL_570159\|2020-04 | ···················· | ···················· | ···················· | ···················· | ···················· | ···················· | |
| K | 2285 | hCoV-19/USA/WA-UW-7013/2020\|EPI_ISL_570161\|2020-04 | ···················· | ···················· | ···················· | ···················· | ···················· | ···················· | |
| K | 2286 | hCoV-19/USA/WA-UW-7022/2020\|EPI_ISL_570163\|2020-04 | ···················· | ···················· | ···················· | ···················· | ···················· | ···················· | |
| K | 2287 | hCoV-19/USA/WA-UW-7032/2020\|EPI_ISL_570164\|2020-04 | ···················· | ···················· | ···················· | ···················· | ···················· | ···················· | |
| K | 2288 | hCoV-19/USA/WA-UW-7040/2020\|EPI_ISL_570165\|2020-04 | ···················· | ···················· | ···················· | ···················· | ···················· | ···················· | |
| K | 2289 | hCoV-19/USA/CT-UW-7070/2020\|EPI_ISL_570166\|2020-04 | ···················· | ···················· | ···················· | ···················· | ···················· | ···················· | |
| K | 2290 | hCoV-19/USA/WA-UW-7093/2020\|EPI_ISL_570167\|2020-04 | ···················· | ···················· | ···················· | ···················· | ···················· | ···················· | |
| K | 2291 | hCoV-19/USA/WA-UW-7128/2020\|EPI_ISL_570170\|2020-04 | ···················· | ···················· | ···················· | ···················· | ···················· | ···················· | |
| K | 2292 | hCoV-19/USA/WA-UW-7160/2020\|EPI_ISL_570172\|2020-04 | ···················· | ···················· | ···················· | ···················· | ···················· | ···················· | |
| K | 2293 | hCoV-19/USA/WA-UW-7164/2020\|EPI_ISL_570173\|2020-04 | ···················· | ···················· | ···················· | ···················· | ···················· | ···················· | |
| K | 2294 | hCoV-19/USA/WA-UW-7236/2020\|EPI_ISL_570175\|2020-04 | ···················· | ···················· | ···················· | ···················· | ···················· | ···················· | |
| K | 2295 | hCoV-19/USA/WA-UW-7237/2020\|EPI_ISL_570176\|2020-04 | ···················· | ···················· | ···················· | ···················· | ···················· | ···················· | |
| K | 2296 | hCoV-19/USA/WA-UW-7242/2020\|EPI_ISL_570177\|2020-04 | ···················· | ···················· | ···················· | ···················· | ···················· | ···················· | |
| K | 2297 | hCoV-19/USA/CT-UW-7257/2020\|EPI_ISL_570178\|2020-04 | ···················· | ···················· | ···················· | ···················· | ···················· | ···················· | |
| K | 2298 | hCoV-19/USA/CT-UW-7260/2020\|EPI_ISL_570179\|2020-04 | ···················· | ···················· | ···················· | ···················· | ···················· | ···················· | |
| K | 2299 | hCoV-19/USA/CT-UW-7265/2020\|EPI_ISL_570180\|2020-04 | ···················· | ···················· | ···················· | ···················· | ···················· | ···················· | |
| K | 2300 | hCoV-19/USA/CT-UW-7266/2020\|EPI_ISL_570181\|2020-04 | ···················· | ···················· | ···················· | ···················· | ···················· | ···················· | |
| K | 2301 | hCoV-19/USA/CT-UW-7270/2020\|EPI_ISL_570182\|2020-04 | ···················· | ···················· | ···················· | ···················· | ···················· | ···················· | |
| K | 2302 | hCoV-19/USA/CT-UW-7285/2020\|EPI_ISL_570184\|2020-04 | ···················· | ···················· | ···················· | ···················· | ···················· | ···················· | |
| K | 2303 | hCoV-19/USA/CT-UW-7290/2020\|EPI_ISL_570185\|2020-04 | ···················· | ···················· | ···················· | ···················· | ···················· | ···················· | |
| K | 2304 | hCoV-19/USA/ID-UW-7291/2020\|EPI_ISL_570186\|2020-04 | ···················· | ···················· | ···················· | ···················· | ···················· | ···················· | |
| K | 2305 | hCoV-19/Australia/VIC13375/2020\|EPI_ISL_563700\|202 | ···················· | ···················· | ···················· | ···················· | ···················· | ···················· | |
| K | 2306 | hCoV-19/Australia/VIC13376/2020\|EPI_ISL_563701\|202 | ···················· | ···················· | ···················· | ···················· | ···················· | ···················· | |
| K | 2307 | hCoV-19/Australia/VIC13377/2020\|EPI_ISL_563702\|202 | ···················· | ···················· | ···················· | ···················· | ···················· | ···················· | |
| K | 2308 | hCoV-19/Australia/VIC13379/2020\|EPI_ISL_563703\|202 | ···················· | ···················· | ···················· | ···················· | ···················· | ···················· | |
| K | 2309 | hCoV-19/Australia/VIC13380/2020\|EPI_ISL_563704\|202 | ···················· | ···················· | ···················· | ···················· | ···················· | ···················· | |
| K | 2310 | hCoV-19/Australia/VIC13382/2020\|EPI_ISL_563705\|202 | ···················· | ···················· | ···················· | ···················· | ···················· | ···················· | |
| K | 2311 | hCoV-19/Australia/VIC13383/2020\|EPI_ISL_563706\|202 | ···················· | ···················· | ···················· | ···················· | ····R··············· | ···················· | |
| K | 2312 | hCoV-19/Australia/VIC13384/2020\|EPI_ISL_563707\|202 | ···················· | ···················· | ···················· | ···················· | ···················· | ···················· | |
| K | 2313 | hCoV-19/Australia/VIC13386/2020\|EPI_ISL_563708\|202 | ···················· | ···················· | ···················· | ···················· | ···················· | ···················· | |
| K | 2315 | hCoV-19/Australia/VIC9556/2020\|EPI_ISL_565600\|2020 | ···················· | ···················· | ···················· | ···················· | ···················· | ···················· | |
| K | 2316 | hCoV-19/Australia/VIC9558/2020\|EPI_ISL_565601\|2020 | ···················· | ···················· | ···················· | ···················· | ···················· | ···················· | |
| K | 2317 | hCoV-19/Australia/VIC9559/2020\|EPI_ISL_565602\|2020 | ···················· | ···················· | ···················· | ···················· | ···················· | ···················· | |
| K | 2318 | hCoV-19/Australia/VIC9562/2020\|EPI_ISL_565603\|2020 | ···················· | ···················· | ···················· | ···················· | ···················· | ···················· | |
| K | 2319 | hCoV-19/Australia/VIC9564/2020\|EPI_ISL_565604\|2020 | ···················· | ···················· | ···················· | ···················· | ···················· | ···················· | |
| K | 2320 | hCoV-19/Australia/VIC9567/2020\|EPI_ISL_565605\|2020 | ···················· | ···················· | ···················· | ···················· | ···················· | ···················· | |
| K | 2321 | hCoV-19/Australia/VIC9570/2020\|EPI_ISL_565606\|2020 | ···················· | ···················· | ···················· | ···················· | ···················· | ···················· | |
| K | 2322 | hCoV-19/Australia/VIC9572/2020\|EPI_ISL_565607\|2020 | ···················· | ···················· | ···················· | ···················· | ···················· | ···················· | |
| K | 2323 | hCoV-19/Australia/VIC9573/2020\|EPI_ISL_565608\|2020 | ···················· | ···················· | ···················· | ···················· | ···················· | ···················· | |
| K | 2324 | hCoV-19/Australia/VIC9574/2020\|EPI_ISL_565609\|2020 | ···················· | ···················· | ···················· | ···················· | ···················· | ···················· | |
| K | 2325 | hCoV-19/Australia/VIC9579/2020\|EPI_ISL_565610\|2020 | ···················· | ···················· | ···················· | ···················· | ···················· | ···················· | |
| K | 2326 | hCoV-19/England/MILK-9C3A11/2020\|EPI_ISL_566920\|20 | ···················· | ···················· | ···················· | ···················· | ···················· | ···················· | |
| K | 2327 | hCoV-19/England/MILK-9C4719/2020\|EPI_ISL_566921\|20 | ···················· | ·A·················· | ···················· | ···················· | ···················· | ···················· | |
| K | 2328 | hCoV-19/England/MILK-9C3DFA/2020\|EPI_ISL_566923\|20 | ···················· | ···················· | ···················· | ···················· | ····T··············· | ···················· | |
| K | 2329 | hCoV-19/England/MILK-9C3FF4/2020\|EPI_ISL_566924\|20 | ···················· | ···················· | ···················· | ···················· | ···················· | ···················· | |
| K | 2330 | hCoV-19/England/MILK-9C3AF3/2020\|EPI_ISL_566926\|20 | ···················· | ···················· | ···················· | ···················· | ···················· | ···················· | |
| K | 2331 | hCoV-19/Wales/MILK-9C44C7/2020\|EPI_ISL_566928\|2020 | ···················· | ···················· | ···················· | ···················· | ···················· | ···················· | |
| K | 2332 | hCoV-19/England/MILK-9C4324/2020\|EPI_ISL_566929\|20 | ···················· | ···················· | ···················· | ···················· | ······T············· | ···················· | |
| K | 2333 | hCoV-19/England/MILK-9C3C93/2020\|EPI_ISL_566930\|20 | ···················· | ···················· | ···················· | ···················· | ···················· | ···················· | |
| K | 2334 | hCoV-19/England/MILK-9C3B3C/2020\|EPI_ISL_566931\|20 | ···················· | ···················· | ···················· | ···················· | ···················· | ···················· | |
| K | 2335 | hCoV-19/England/MILK-9C3422/2020\|EPI_ISL_566933\|20 | ···················· | ···················· | ···················· | ···················· | ···················· | ···················· | |
| K | 2336 | hCoV-19/England/MILK-9C4078/2020\|EPI_ISL_566934\|20 | ···················· | ···················· | ···················· | ···················· | ···················· | ···················· | |
| K | 2337 | hCoV-19/England/MILK-9C30F1/2020\|EPI_ISL_566935\|20 | ···················· | ···················· | ···················· | ···················· | ···················· | ···················· | |
| K | 2338 | hCoV-19/England/MILK-9C357A/2020\|EPI_ISL_566937\|20 | ···················· | ···················· | ···················· | ···················· | ···················· | ···················· | |
| K | 2339 | hCoV-19/England/MILK-9C445E/2020\|EPI_ISL_566938\|20 | ···················· | ·A·················· | ···················· | ···················· | ···················· | ···A················ | |
| K | 2340 | hCoV-19/England/MILK-9C4193/2020\|EPI_ISL_566939\|20 | ···················· | ···················· | ···················· | ···················· | ···················· | ···················· | |
| K | 2341 | hCoV-19/England/MILK-9C3185/2020\|EPI_ISL_566940\|20 | ···················· | ·A·················· | ···················· | ···················· | ···················· | ···A················ | |
| K | 2342 | hCoV-19/England/MILK-9C470A/2020\|EPI_ISL_566941\|20 | ···················· | ···················· | ···················· | ···················· | ···················· | ···················· | |
| K | 2343 | hCoV-19/England/QEUH-9C8FC2/2020\|EPI_ISL_567206\|20 | ···················· | ···················· | ···················· | ···················· | ···················· | ···················· | |
| K | 2344 | hCoV-19/England/QEUH-9C9037/2020\|EPI_ISL_567207\|20 | ···················· | ···················· | ···················· | ···················· | ···················· | ···················· | |
| K | 2345 | hCoV-19/England/QEUH-9C845A/2020\|EPI_ISL_567208\|20 | ···················· | ···················· | ···················· | ···················· | ···················· | ···················· | |
| K | 2346 | hCoV-19/England/QEUH-9C8B82/2020\|EPI_ISL_567209\|20 | ···················· | ···················· | ···················· | ···················· | ···················· | ···················· | |
| K | 2347 | hCoV-19/England/QEUH-9C833F/2020\|EPI_ISL_567215\|20 | ···················· | ···················· | ···················· | ···················· | ···················· | ···················· | |
| K | 2348 | hCoV-19/England/QEUH-9C8B73/2020\|EPI_ISL_567216\|20 | ···················· | ···················· | ···················· | ···················· | ···················· | ···················· | |
| K | 2349 | hCoV-19/England/QEUH-9C835D/2020\|EPI_ISL_567239\|20 | ···················· | ···················· | ···················· | ···················· | ···················· | ···················· | |
| K | 2350 | hCoV-19/England/QEUH-9C8126/2020\|EPI_ISL_567240\|20 | ···················· | ···················· | ···················· | ···················· | ···················· | ···················· | |
| K | 2351 | hCoV-19/England/QEUH-9C86CD/2020\|EPI_ISL_567242\|20 | ···················· | ···················· | ···················· | ···················· | ···················· | ···················· | |

| | | | | | | | |
|---|---|---|---|---|---|---|---|
| K | 2352 hCoV-19/England/QEUH-9C81AE/2020\|EPI_ISL_567243\|20 | ···················· | ···················· | ····················· | ···················· | ···················· | ···················· |
| K | 2353 hCoV-19/England/QEUH-9C891F/2020\|EPI_ISL_567245\|20 | ···················· | ···················· | ····················· | ···················· | ···················· | ···················· |
| K | 2354 hCoV-19/England/QEUH-9C80A1/2020\|EPI_ISL_567246\|20 | ···················· | ···················· | ····················· | ···················· | ···················· | ···················· |
| K | 2355 hCoV-19/England/QEUH-9C89B5/2020\|EPI_ISL_567247\|20 | ···················· | ·A·················· | ····················· | ···················· | ···················· | ···················· |
| K | 2356 hCoV-19/England/QEUH-9C8654/2020\|EPI_ISL_567248\|20 | ···················· | ···················· | ····················· | ···················· | ···················· | ···················· |
| K | 2357 hCoV-19/England/QEUH-9C7E6C/2020\|EPI_ISL_567249\|20 | ···················· | ···················· | ····················· | ···················· | ···················· | ···················· |
| K | 2358 hCoV-19/England/QEUH-9C886D/2020\|EPI_ISL_567250\|20 | ···················· | ···················· | ····················· | ···················· | ···················· | ···················· |
| K | 2359 hCoV-19/England/QEUH-9C7D7E/2020\|EPI_ISL_567251\|20 | ···················· | ·A·················· | ····················· | ···················· | ···················· | ···A················ |
| K | 2360 hCoV-19/England/QEUH-9C8DAA/2020\|EPI_ISL_567252\|20 | ···················· | ···················· | ····················· | ···················· | ···················· | ···················· |
| K | 2361 hCoV-19/England/QEUH-9C88C7/2020\|EPI_ISL_567253\|20 | ···················· | ···················· | ····················· | ···················· | ······T············· | ···················· |
| K | 2362 hCoV-19/England/QEUH-9C81CC/2020\|EPI_ISL_567254\|20 | ···················· | ···················· | ····················· | ···················· | ···················· | ···················· |
| K | 2363 hCoV-19/Wales/QEUH-9C87AC/2020\|EPI_ISL_567255\|2020 | ···················· | ···················· | ····················· | ···················· | ···················· | ···················· |
| K | 2364 hCoV-19/England/QEUH-9C826F/2020\|EPI_ISL_567256\|20 | ···················· | ···················· | ····················· | ···················· | ···················· | ···················· |
| K | 2365 hCoV-19/England/QEUH-9C8065/2020\|EPI_ISL_567257\|20 | ···················· | ···················· | ····················· | ···················· | ···················· | ···················· |
| K | 2366 hCoV-19/England/QEUH-9C8038/2020\|EPI_ISL_567258\|20 | ···················· | ·A·················· | ····················· | ···················· | ···················· | ···················· |
| K | 2367 hCoV-19/England/QEUH-9C8627/2020\|EPI_ISL_567259\|20 | ···················· | ···················· | ····················· | ···················· | ···················· | ···················· |
| K | 2368 hCoV-19/England/QEUH-9C8575/2020\|EPI_ISL_567261\|20 | ···················· | ···················· | ····················· | ···················· | ···················· | ···················· |
| K | 2369 hCoV-19/England/QEUH-9C895B/2020\|EPI_ISL_567262\|20 | ···················· | ···················· | ····················· | ···················· | ···················· | ···················· |
| K | 2370 hCoV-19/England/QEUH-9C8A67/2020\|EPI_ISL_567264\|20 | ···················· | ···················· | ····················· | ···················· | ···················· | ···················· |
| K | 2371 hCoV-19/England/QEUH-9C8135/2020\|EPI_ISL_567266\|20 | ···················· | ·A·················· | ····················· | ···················· | ···················· | ···················· |
| K | 2372 hCoV-19/England/QEUH-9C80ED/2020\|EPI_ISL_567267\|20 | ···················· | ···················· | ····················· | ···················· | ···················· | ···················· |
| K | 2373 hCoV-19/England/QEUH-9C86AF/2020\|EPI_ISL_567268\|20 | ···················· | ···················· | ····················· | ···················· | ···················· | ···················· |
| K | 2374 hCoV-19/England/QEUH-9C9459/2020\|EPI_ISL_567269\|20 | ···················· | ···················· | ····················· | ···················· | ···················· | ···················· |
| K | 2375 hCoV-19/England/QEUH-9C990F/2020\|EPI_ISL_567270\|20 | ···················· | ···················· | ····················· | ···················· | ···················· | ···················· |
| K | 2376 hCoV-19/England/QEUH-9C9996/2020\|EPI_ISL_567271\|20 | ···················· | ···················· | ····················· | ···················· | ···················· | ···················· |
| K | 2377 hCoV-19/England/QEUH-9CA528/2020\|EPI_ISL_567272\|20 | ···················· | ···················· | ····················· | ···················· | ···················· | ···················· |
| K | 2378 hCoV-19/England/QEUH-9C9592/2020\|EPI_ISL_567273\|20 | ···················· | ···················· | ····················· | ···················· | ···················· | ···················· |
| K | 2379 hCoV-19/Wales/QEUH-9C9EA6/2020\|EPI_ISL_567274\|2020 | ···················· | ···················· | ····················· | ···················· | ···················· | ···················· |
| K | 2380 hCoV-19/England/QEUH-9C95FC/2020\|EPI_ISL_567275\|20 | ···················· | ···················· | ····················· | ···················· | ···················· | ···················· |
| K | 2381 hCoV-19/England/QEUH-9CA652/2020\|EPI_ISL_567276\|20 | ···················· | ···················· | ····················· | ···················· | ···················· | ···················· |
| K | 2382 hCoV-19/England/QEUH-9C941D/2020\|EPI_ISL_567277\|20 | ···················· | ···················· | ····················· | ···················· | ···················· | ···················· |
| K | 2383 hCoV-19/England/QEUH-9C940E/2020\|EPI_ISL_567279\|20 | ···················· | ···················· | ····················· | ···················· | ···················· | ···················· |
| K | 2384 hCoV-19/England/QEUH-9CA704/2020\|EPI_ISL_567281\|20 | ···················· | ···················· | ····················· | ···················· | ···················· | ···················· |
| K | 2385 hCoV-19/England/QEUH-9CA31F/2020\|EPI_ISL_567282\|20 | ···················· | ···················· | ····················· | ···················· | ···················· | ···················· |
| K | 2386 hCoV-19/England/QEUH-9C97AB/2020\|EPI_ISL_567283\|20 | ···················· | ···················· | ····················· | ···················· | ······T············· | ···················· |
| K | 2387 hCoV-19/England/QEUH-9C9477/2020\|EPI_ISL_567284\|20 | ···················· | ···················· | ·········T··········· | ···················· | ······T············· | ···················· |
| K | 2388 hCoV-19/England/QEUH-9CA3F1/2020\|EPI_ISL_567285\|20 | ···················· | ···················· | ····················· | ···················· | ···················· | ···················· |
| K | 2389 hCoV-19/England/QEUH-9C950B/2020\|EPI_ISL_567286\|20 | ···················· | ···················· | ····················· | ···················· | ···················· | ···················· |
| K | 2390 hCoV-19/England/QEUH-9C9750/2020\|EPI_ISL_567287\|20 | ···················· | ···················· | ····················· | ···················· | ···················· | ···················· |
| K | 2391 hCoV-19/England/QEUH-9C9617/2020\|EPI_ISL_567288\|20 | ···················· | ···················· | ····················· | ···················· | ···················· | ···················· |
| K | 2392 hCoV-19/England/QEUH-9CA1BB/2020\|EPI_ISL_567290\|20 | ···················· | ····A··············· | ····················· | ···················· | ···················· | ···················· |
| K | 2393 hCoV-19/England/QEUH-9C9C06/2020\|EPI_ISL_567291\|20 | ···················· | ···················· | ····················· | ···················· | ···················· | ···················· |
| K | 2394 hCoV-19/England/QEUH-9C9C7F/2020\|EPI_ISL_567292\|20 | ···················· | ···················· | ····················· | ···················· | ···················· | ···················· |
| K | 2395 hCoV-19/Wales/QEUH-9CA19D/2020\|EPI_ISL_567293\|2020 | ···················· | ···················· | ····················· | ···················· | ···················· | ···················· |
| K | 2396 hCoV-19/England/QEUH-9CA142/2020\|EPI_ISL_567294\|20 | ···················· | ···················· | ····················· | ···················· | ······T············· | ···················· |
| K | 2397 hCoV-19/England/QEUH-9C9635/2020\|EPI_ISL_567297\|20 | ···················· | ···················· | ····················· | ···················· | ···················· | ···················· |
| K | 2398 hCoV-19/Wales/QEUH-9C9820/2020\|EPI_ISL_567299\|2020 | ···················· | ···················· | ····················· | ···················· | ···················· | ···················· |
| K | 2399 hCoV-19/Peru/LIM-UPCH-0028/2020\|EPI_ISL_568510\|202 | ···················· | ···················· | ····················· | ···················· | ···················· | ···················· |
| K | 2400 hCoV-19/Peru/LIM-UPCH-0054/2020\|EPI_ISL_568511\|202 | ···················· | ···················· | ····················· | ···················· | ···················· | ···················· |
| K | 2401 hCoV-19/Peru/LIM-UPCH-0077/2020\|EPI_ISL_568512\|202 | ···················· | ···················· | ····················· | ···················· | ···················· | ···················· |
| K | 2402 hCoV-19/Peru/LIM-UPCH-0116/2020\|EPI_ISL_568513\|202 | ···················· | ···················· | ····················· | ···················· | ···················· | ···················· |
| K | 2403 hCoV-19/Peru/LIM-UPCH-0117/2020\|EPI_ISL_568514\|202 | ···················· | ···················· | ····················· | ···················· | ···················· | ···················· |
| K | 2404 hCoV-19/Peru/JUN-UPCH-0118/2020\|EPI_ISL_568515\|202 | ···················· | ···················· | ····················· | ···················· | ···················· | ···················· |
| K | 2405 hCoV-19/Peru/LIM-UPCH-0119/2020\|EPI_ISL_568516\|202 | ···················· | ···················· | ····················· | ···················· | ···················· | ···················· |
| K | 2406 hCoV-19/Peru/LIM-UPCH-0120/2020\|EPI_ISL_568517\|202 | ···················· | ···················· | ····················· | ···················· | ···················· | ···················· |
| K | 2407 hCoV-19/Peru/LIM-UPCH-0122/2020\|EPI_ISL_568518\|202 | ···················· | ···················· | ····················· | ···················· | ···················· | ···················· |
| K | 2408 hCoV-19/Peru/LIM-UPCH-0123/2020\|EPI_ISL_568519\|202 | ···················· | ···················· | ····················· | ···················· | ···················· | ···················· |
| K | 2409 hCoV-19/Peru/LIM-UPCH-0124/2020\|EPI_ISL_568520\|202 | ···················· | ···················· | ····················· | ···················· | ···················· | ···················· |
| K | 2410 hCoV-19/Peru/LIM-UPCH-0125/2020\|EPI_ISL_568521\|202 | ···················· | ···················· | ····················· | ···················· | ···················· | ···················· |
| K | 2411 hCoV-19/Peru/LIM-UPCH-0126/2020\|EPI_ISL_568522\|202 | ···················· | ···················· | ····················· | ···················· | ···················· | ···················· |
| K | 2412 hCoV-19/Peru/LIM-UPCH-0127/2020\|EPI_ISL_568523\|202 | ···················· | ···················· | ····················· | ···················· | ···················· | ···················· |
| K | 2413 hCoV-19/Peru/LIM-UPCH-0128/2020\|EPI_ISL_568524\|202 | ···················· | ···················· | ····················· | ···················· | ···················· | ···················· |
| K | 2414 hCoV-19/Peru/LIM-UPCH-0129/2020\|EPI_ISL_568525\|202 | ···················· | ···················· | ····················· | ···················· | ···················· | ···················· |
| K | 2415 hCoV-19/Peru/LIM-UPCH-0143/2020\|EPI_ISL_568537\|202 | ···················· | ···················· | ····················· | ···················· | ···················· | ···················· |
| K | 2416 hCoV-19/Peru/LIM-UPCH-0144/2020\|EPI_ISL_568538\|202 | ···················· | ···················· | ····················· | ···················· | ···················· | ···················· |
| K | 2417 hCoV-19/Peru/LIM-UPCH-0145/2020\|EPI_ISL_568539\|202 | ···················· | ···················· | ····················· | ···················· | ···················· | ···················· |
| K | 2418 hCoV-19/Peru/LIM-UPCH-0150/2020\|EPI_ISL_568544\|202 | ···················· | ···················· | ····················· | ···················· | ···················· | ···················· |
| K | 2419 hCoV-19/Peru/LIM-UPCH-0151/2020\|EPI_ISL_568545\|202 | ···················· | ···················· | ····················· | ···················· | ···················· | ···················· |
| K | 2420 hCoV-19/Peru/LIM-UPCH-0152/2020\|EPI_ISL_568546\|202 | ···················· | ···················· | ····················· | ···················· | ···················· | ···················· |
| K | 2421 hCoV-19/Peru/LIM-UPCH-0154/2020\|EPI_ISL_568547\|202 | ···················· | ···················· | ····················· | ···················· | ···················· | ···················· |
| K | 2422 hCoV-19/Japan/NGY-NNH-015/2020\|EPI_ISL_568570\|2020 | ···················· | ···················· | ····················· | ···················· | ···················· | ···················· |
| K | 2423 hCoV-19/Japan/NGY-NNH-016/2020\|EPI_ISL_568571\|2020 | ···················· | ···················· | ····················· | ···················· | ···················· | ···················· |
| K | 2424 hCoV-19/Japan/NGY-NNH-017/2020\|EPI_ISL_568572\|2020 | ···················· | ···················· | ····················· | ···················· | ···················· | ···················· |
| K | 2425 hCoV-19/Japan/NGY-NNH-018/2020\|EPI_ISL_568573\|2020 | ···················· | ···················· | ····················· | ···················· | ···················· | ···················· |
| K | 2426 hCoV-19/Japan/NGY-NNH-019/2020\|EPI_ISL_568574\|2020 | ···················· | ···················· | ····················· | ···················· | ···················· | ···················· |
| K | 2427 hCoV-19/Japan/NGY-NNH-020/2020\|EPI_ISL_568575\|2020 | ···················· | ···················· | ····················· | ···················· | ···················· | ···················· |
| K | 2428 hCoV-19/Japan/NGY-NNH-021/2020\|EPI_ISL_568576\|2020 | ···················· | ···················· | ····················· | ···················· | ···················· | ···················· |
| K | 2429 hCoV-19/Czech Republic/NRL-8631/2020\|EPI_ISL_56857 | ···················· | ·A·················· | ····················· | ···················· | ···················· | ···················· |
| K | 2430 hCoV-19/Italy/LOM-INMI-10734/2020\|EPI_ISL_568579\|2 | ···················· | ···················· | ····················· | ···················· | ···················· | ···················· |
| K | 2431 hCoV-19/Thailand/Bangkok-CONI-0215/2020\|EPI_ISL_56 | ···················· | ···················· | ····················· | ···················· | ···················· | ···················· |

| | | | | | | | | |
|---|---|---|---|---|---|---|---|---|
| K | 2432 | hCoV-19/Thailand/Bangkok-CONI-0216/2020\|EPI_ISL_56 | .................... | .................... | ..................... | .................... | .................... | .................... |
| K | 2433 | hCoV-19/Thailand/Bangkok-CONI-0217/2020\|EPI_ISL_56 | .................... | .................... | ..................... | .................... | .................... | .................... |
| K | 2434 | hCoV-19/Thailand/Bangkok-CONI-0219/2020\|EPI_ISL_56 | .................... | .................... | ..................... | .................... | .................... | .................... |
| K | 2435 | hCoV-19/Thailand/Bangkok-CONI-0220/2020\|EPI_ISL_56 | .................... | .................... | ..................... | .................... | .................... | .................... |
| K | 2436 | hCoV-19/Thailand/Bangkok-CONI-0221/2020\|EPI_ISL_56 | .................... | .................... | ..................... | .................... | .................... | .................... |
| K | 2437 | hCoV-19/Thailand/Bangkok-CONI-0222/2020\|EPI_ISL_56 | .................... | .................... | ..................... | .................... | .................... | .................... |
| K | 2438 | hCoV-19/USA/FL-BPHL-1489/2020\|EPI_ISL_568587\|2020- | .................... | .................... | ..................... | .................... | .................... | .................... |
| K | 2439 | hCoV-19/USA/FL-BPHL-1492/2020\|EPI_ISL_568590\|2020- | .................... | .................... | ..................... | .................... | .................... | .................... |
| K | 2440 | hCoV-19/USA/FL-BPHL-1493/2020\|EPI_ISL_568591\|2020- | .................... | .................... | ..................... | .................... | .................... | .................... |
| K | 2441 | hCoV-19/USA/FL-BPHL-1497/2020\|EPI_ISL_568595\|2020- | .................... | .................... | ..................... | .................... | .................... | .................... |
| K | 2442 | hCoV-19/USA/FL-BPHL-1499/2020\|EPI_ISL_568597\|2020- | .................... | .................... | ..................... | .................... | .................... | .................... |
| K | 2443 | hCoV-19/USA/FL-BPHL-1500/2020\|EPI_ISL_568598\|2020- | .................... | .................... | ..................... | .................... | .................... | .................... |
| K | 2444 | hCoV-19/USA/FL-BPHL-1501/2020\|EPI_ISL_568599\|2020- | .................... | .................... | ..................... | .................... | .................... | .................... |
| K | 2445 | hCoV-19/France/PAC-IHU-1505/2020\|EPI_ISL_569181\|20 | .................... | .................... | ..................... | .................... | .................... | .................... |
| K | 2446 | hCoV-19/France/PAC-IHU-1509/2020\|EPI_ISL_569182\|20 | .................... | .................... | ..................... | .................... | .................... | .................... |
| K | 2447 | hCoV-19/France/PAC-IHU-1511/2020\|EPI_ISL_569183\|20 | .................... | .................... | ..................... | .................... | .................... | .................... |
| K | 2448 | hCoV-19/France/PAC-IHU-1513/2020\|EPI_ISL_569184\|20 | .................... | ....A............... | ..................... | .................... | .................... | .................... |
| K | 2449 | hCoV-19/France/PAC-IHU-1519/2020\|EPI_ISL_569185\|20 | .................... | .................... | ..................... | .................... | .................... | .................... |
| K | 2450 | hCoV-19/France/PAC-IHU-1521/2020\|EPI_ISL_569186\|20 | .................... | .................... | ..................... | .................... | .................... | .................... |
| K | 2451 | hCoV-19/Russia/OMS-ORINFI-5286S/2020\|EPI_ISL_56981 | .................... | .................... | ..................... | .................... | .................... | .................... |
| K | 2452 | hCoV-19/Russia/OMS-ORINFI-3970S/2020\|EPI_ISL_56981 | .................... | .................... | ..................... | .................... | .................... | .................... |
| K | 2453 | hCoV-19/Russia/OMS-ORINFI-3873S/2020\|EPI_ISL_56981 | .................... | .................... | ..................... | .................... | .................... | .................... |
| K | 2454 | hCoV-19/Russia/OMS-ORINFI-4828S/2020\|EPI_ISL_56981 | .................... | .................... | ..................... | .................... | .................... | .................... |
| K | 2455 | hCoV-19/Russia/OMS-ORINFI-47S/2020\|EPI_ISL_569817\| | .................... | .................... | ..................... | .................... | .................... | .................... |
| K | 2456 | hCoV-19/Russia/OMS-ORINFI-120S/2020\|EPI_ISL_569818 | .................... | .................... | ..................... | .................... | .................... | .................... |
| K | 2457 | hCoV-19/Russia/OMS-ORINFI-1630S/2020\|EPI_ISL_56981 | .................... | .................... | ..................... | .................... | .................... | .................... |
| K | 2458 | hCoV-19/Russia/OMS-ORINFI-4791S/2020\|EPI_ISL_56982 | .................... | .................... | ..................... | .................... | .................... | .................... |
| K | 2459 | hCoV-19/Russia/OMS-ORINFI-3S/2020\|EPI_ISL_569821\|2 | .................... | .................... | ..................... | .................... | .................... | .................... |
| K | 2460 | hCoV-19/Russia/OMS-ORINFI-78S/2020\|EPI_ISL_569822\| | .................... | .................... | ..................... | .................... | .................... | .................... |
| K | 2461 | hCoV-19/Russia/OMS-ORINFI-5195S/2020\|EPI_ISL_56982 | .................... | .................... | ..................... | .................... | .................... | .................... |
| K | 2462 | hCoV-19/Russia/OMS-ORINFI-6957S/2020\|EPI_ISL_56982 | .................... | .................... | ..................... | .................... | .................... | .................... |
| K | 2463 | hCoV-19/Russia/OMS-ORINFI-55S/2020\|EPI_ISL_569825\| | .................... | .................... | ..................... | .................... | .................... | .................... |
| K | 2464 | hCoV-19/Russia/OMS-ORINFI-2535S/2020\|EPI_ISL_56982 | .................... | .................... | ..................... | .................... | .................... | .................... |
| K | 2465 | hCoV-19/Russia/OMS-ORINFI-1631S/2020\|EPI_ISL_56982 | .................... | .................... | ..................... | .................... | .................... | .................... |
| K | 2466 | hCoV-19/Russia/OMS-ORINFI-2112S/2020\|EPI_ISL_56982 | .................... | .................... | ..................... | .................... | .................... | .................... |
| K | 2467 | hCoV-19/Russia/OMS-ORINFI-4814S/2020\|EPI_ISL_56982 | .................... | .................... | ..................... | .................... | .................... | .................... |
| K | 2468 | hCoV-19/Russia/OMS-ORINFI-5199S/2020\|EPI_ISL_56983 | .................... | .................... | ..................... | .................... | .................... | .................... |
| K | 2469 | hCoV-19/Russia/OMS-ORINFI-8436S/2020\|EPI_ISL_56983 | .................... | .................... | ..................... | .................... | .................... | .................... |
| K | 2470 | hCoV-19/Russia/OMS-ORINFI-56S/2020\|EPI_ISL_569832\| | .................... | .................... | ..................... | .................... | .................... | .................... |
| K | 2471 | hCoV-19/Russia/OMS-ORINFI-1632S/2020\|EPI_ISL_56983 | .................... | .................... | ..................... | .................... | .................... | .................... |
| K | 2472 | hCoV-19/Russia/OMS-ORINFI-3961S/2020\|EPI_ISL_56983 | .................... | .................... | ..................... | .................... | .................... | .................... |
| K | 2473 | hCoV-19/Russia/OMS-ORINFI-2427S/2020\|EPI_ISL_56983 | .................... | .................... | ..................... | .................... | .................... | .................... |
| K | 2474 | hCoV-19/Russia/OMS-ORINFI-1141S/2020\|EPI_ISL_56983 | .................... | .................... | ..................... | .................... | .................... | .................... |
| K | 2475 | hCoV-19/Russia/OMS-ORINFI-106S/2020\|EPI_ISL_569837 | .................... | .................... | ..................... | .................... | .................... | .................... |
| K | 2476 | hCoV-19/Russia/OMS-ORINFI-5204S/2020\|EPI_ISL_56983 | .................... | .................... | ..................... | .................... | .................... | .................... |
| K | 2477 | hCoV-19/Russia/OMS-ORINFI-8474S/2020\|EPI_ISL_56983 | .................... | .................... | ..................... | .................... | .................... | .................... |
| K | 2478 | hCoV-19/Russia/OMS-ORINFI-57S/2020\|EPI_ISL_569840\| | .................... | .................... | ..................... | .................... | .................... | .................... |
| K | 2479 | hCoV-19/Russia/OMS-ORINFI-2887S/2020\|EPI_ISL_56984 | .................... | .................... | ..................... | .................... | .................... | .................... |
| K | 2480 | hCoV-19/Russia/OMS-ORINFI-1639S/2020\|EPI_ISL_56984 | .................... | .................... | ..................... | .................... | .................... | .................... |
| K | 2481 | hCoV-19/Russia/OMS-ORINFI-9661S/2020\|EPI_ISL_56984 | .................... | .................... | ..................... | .................... | .................... | .................... |
| K | 2482 | hCoV-19/Russia/OMS-ORINFI-7593S/2020\|EPI_ISL_56984 | .................... | .................... | ..................... | .................... | .................... | .................... |
| K | 2483 | hCoV-19/Russia/OMS-ORINFI-4818S/2020\|EPI_ISL_56984 | .................... | .................... | ..................... | .................... | .................... | .................... |
| K | 2484 | hCoV-19/Russia/OMS-ORINFI-5208S/2020\|EPI_ISL_56984 | .................... | .................... | ..................... | .................... | .................... | .................... |
| K | 2485 | hCoV-19/Russia/OMS-ORINFI-8499S/2020\|EPI_ISL_56984 | .................... | .................... | ..................... | .................... | .................... | .................... |
| K | 2486 | hCoV-19/Russia/OMS-ORINFI-58S/2020\|EPI_ISL_569848\| | .................... | .................... | ..................... | .................... | .................... | .................... |
| K | 2487 | hCoV-19/Russia/OMS-ORINFI-3401S/2020\|EPI_ISL_56984 | .................... | .................... | ..................... | .................... | .................... | .................... |
| K | 2488 | hCoV-19/Russia/OMS-ORINFI-3951S/2020\|EPI_ISL_56985 | .................... | .................... | ..................... | .................... | .................... | .................... |
| K | 2489 | hCoV-19/Russia/OMS-ORINFI-7598S/2020\|EPI_ISL_56985 | .................... | .................... | ..................... | .................... | .................... | .................... |
| K | 2490 | hCoV-19/Russia/OMS-ORINFI-9975S/2020\|EPI_ISL_56985 | .................... | .................... | ..................... | .................... | .................... | .................... |
| K | 2491 | hCoV-19/Russia/OMS-ORINFI-4368S/2020\|EPI_ISL_56985 | .................... | .................... | ..................... | .................... | .................... | .................... |
| K | 2492 | hCoV-19/Russia/OMS-ORINFI-4890S/2020\|EPI_ISL_56985 | .................... | .................... | ..................... | .................... | .................... | .................... |
| K | 2493 | hCoV-19/Russia/OMS-ORINFI-60S/2020\|EPI_ISL_569855\| | .................... | .................... | ..................... | .................... | .................... | .................... |
| K | 2494 | hCoV-19/Russia/OMS-ORINFI-9060S/2020\|EPI_ISL_56985 | .................... | .................... | ..................... | .................... | .................... | .................... |
| K | 2495 | hCoV-19/Italy/PIE-SLL-MS52/2020\|EPI_ISL_569868\|202 | .................... | .................... | ..................... | .................... | .................... | .................... |
| K | 2496 | hCoV-19/Italy/PIE-SLL-MS53/2020\|EPI_ISL_569869\|202 | .................... | .................... | ..................... | .................... | .................... | .................... |
| K | 2497 | hCoV-19/Italy/PIE-SLL-MS59/2020\|EPI_ISL_569875\|202 | .................... | .................... | ..................... | .................... | .................... | .................... |
| K | 2498 | hCoV-19/Italy/PIE-SLL-MS61/2020\|EPI_ISL_569876\|202 | .................... | .................... | ..................... | .................... | .................... | .................... |
| K | 2499 | hCoV-19/Italy/PIE-SLL-MS62/2020\|EPI_ISL_569877\|202 | .................... | .................... | ..................... | .................... | .................... | .................... |
| K | 2500 | hCoV-19/Italy/PIE-SLL-MS63/2020\|EPI_ISL_569878\|202 | .................... | .................... | ..................... | .................... | .................... | .................... |
| K | 2501 | hCoV-19/USA/WA-UW-7106/2020\|EPI_ISL_570168\|2020-04 | .................... | .................... | ..................... | .................... | .................... | .................... |
| K | 2502 | hCoV-19/USA/WA-UW-7115/2020\|EPI_ISL_570169\|2020-04 | .................... | .................... | ..................... | .................... | .................... | .................... |
| K | 2503 | hCoV-19/USA/ID-UW-7306/2020\|EPI_ISL_570187\|2020-04 | .................... | .................... | ..................... | .................... | .................... | .................... |
| K | 2504 | hCoV-19/USA/ID-UW-7308/2020\|EPI_ISL_570188\|2020-04 | .................... | .................... | ..................... | .................... | .................... | .................... |
| K | 2505 | hCoV-19/USA/WA-UW-7671/2020\|EPI_ISL_570190\|2020-04 | .................... | .................... | ..................... | .................... | .................... | .................... |
| K | 2506 | hCoV-19/USA/WA-UW-7675/2020\|EPI_ISL_570191\|2020-04 | .................... | .................... | ..................... | .................... | .................... | .................... |
| K | 2507 | hCoV-19/USA/WA-UW-7763/2020\|EPI_ISL_570192\|2020-04 | .................... | .................... | ..................... | .................... | .................... | .................... |
| K | 2508 | hCoV-19/USA/WA-UW-7776/2020\|EPI_ISL_570193\|2020-04 | .................... | .................... | ..................... | .................... | .................... | .................... |
| K | 2509 | hCoV-19/USA/WA-UW-7777/2020\|EPI_ISL_570194\|2020-04 | .................... | .................... | ..................... | .................... | .................... | .................... |
| K | 2510 | hCoV-19/USA/WA-UW-8008/2020\|EPI_ISL_570195\|2020-04 | .................... | .................... | ..................... | .................... | .................... | .................... |
| K | 2511 | hCoV-19/USA/WA-UW-8012/2020\|EPI_ISL_570196\|2020-04 | .................... | .................... | ..................... | .................... | .................... | .................... |

K 2512 hCoV-19/USA/WA-UW-8065/2020|EPI_ISL_570197|2020-04 ···················· ···················· ···················· ···················· ···················· ····················
K 2513 hCoV-19/USA/WA-UW-8146/2020|EPI_ISL_570198|2020-04 ···················· ···················· ···················· ···················· ···················· ····················
K 2514 hCoV-19/USA/WA-UW-8158/2020|EPI_ISL_570199|2020-04 ···················· ···················· ···················· ···················· ···················· ····················
K 2515 hCoV-19/USA/WA-UW-12633/2020|EPI_ISL_570818|2020-0 ···················· ···················· ···················· ···················· ···················· ····················
K 2516 hCoV-19/USA/WA-UW-12771/2020|EPI_ISL_570819|2020-0 ···················· ·A·················· ···················· ···················· ···················· ····················
K 2517 hCoV-19/USA/WA-UW-21969/2020|EPI_ISL_570820|2020-0 ···················· ···················· ···················· ···················· ···················· ····················
K 2518 hCoV-19/USA/WA-UW-19536/2020|EPI_ISL_570821|2020-0 ···················· ···················· ···················· ···················· ···················· ····················
K 2519 hCoV-19/USA/WA-UW-21712/2020|EPI_ISL_570822|2020-0 ···················· ···················· ···················· ···················· ···················· ····················
K 2520 hCoV-19/USA/WA-UW-21995/2020|EPI_ISL_570823|2020-0 ···················· ···················· ···················· ···················· ···················· ····················
K 2521 hCoV-19/USA/WA-UW-12632/2020|EPI_ISL_570824|2020-0 ···················· ·A·················· ···················· ···················· ···················· ····················
K 2522 hCoV-19/USA/WA-UW-12657/2020|EPI_ISL_570825|2020-0 ···················· ···················· ···················· ···················· ···················· ····················
K 2523 hCoV-19/USA/WA-UW-21747/2020|EPI_ISL_570826|2020-0 ···················· ···················· ···················· ···················· ···················· ····················
K 2524 hCoV-19/USA/WA-UW-21764/2020|EPI_ISL_570827|2020-0 ···················· ···················· ···················· ···················· ···················· ····················
K 2525 hCoV-19/USA/WA-UW-22020/2020|EPI_ISL_570828|2020-0 ···················· ···················· ···················· ···················· ···················· ····················
K 2526 hCoV-19/USA/WA-UW-22027/2020|EPI_ISL_570829|2020-0 ···················· ···················· ···················· ···················· ···················· ····················
K 2527 hCoV-19/USA/WA-UW-22028/2020|EPI_ISL_570830|2020-0 ···················· ···················· ···················· ···················· ···················· ····················
K 2528 hCoV-19/USA/WA-UW-13090/2020|EPI_ISL_570831|2020-0 ···················· ···················· ···················· ···················· ···················· ····················
K 2529 hCoV-19/USA/WA-UW-21695/2020|EPI_ISL_570832|2020-0 ···················· ·A·················· ···················· ···················· ···················· ····················
K 2530 hCoV-19/USA/WA-UW-12601/2020|EPI_ISL_570833|2020-0 ···················· ·A·················· ···················· ···················· ···················· ····················
K 2531 hCoV-19/USA/WA-UW-13102/2020|EPI_ISL_570834|2020-0 ···················· ···················· ···················· ···················· ···················· ····················
K 2532 hCoV-19/USA/WA-UW-19426/2020|EPI_ISL_570835|2020-0 ···················· ·A·················· ···················· ···················· ···················· ····················
K 2533 hCoV-19/USA/WA-UW-12935/2020|EPI_ISL_570836|2020-0 ···················· ···················· ···················· ···················· ···················· ····················
K 2534 hCoV-19/USA/WA-UW-13016/2020|EPI_ISL_570837|2020-0 ···················· ·A·················· ···················· ···················· ···················· ····················
K 2535 hCoV-19/USA/WA-UW-22017/2020|EPI_ISL_570838|2020-0 ···················· ···················· ···················· ···················· ···················· ····················
K 2536 hCoV-19/USA/WA-UW-21757/2020|EPI_ISL_570839|2020-0 ···················· ···················· ···················· ···················· ···················· ····················
K 2537 hCoV-19/USA/WA-UW-21726/2020|EPI_ISL_570855|2020-0 ···················· ···················· ···················· ···················· ···················· ····················
K 2538 hCoV-19/USA/WA-UW-21789/2020|EPI_ISL_570856|2020-0 ···················· ···················· ···················· ···················· ···················· ····················
K 2539 hCoV-19/USA/WA-UW-22009/2020|EPI_ISL_570857|2020-0 ···················· ···················· ···················· ···················· ···················· ····················
K 2540 hCoV-19/USA/WA-UW-12772/2020|EPI_ISL_570858|2020-0 ···················· ·A·················· ···················· ···················· ···················· ····················
K 2541 hCoV-19/USA/WA-UW-12627/2020|EPI_ISL_570880|2020-0 ···················· ···················· ···················· ···················· ···················· ····················
K 2542 hCoV-19/USA/WA-UW-12634/2020|EPI_ISL_570881|2020-0 ···················· ···················· ···················· ···················· ···················· ····················
K 2543 hCoV-19/USA/WA-UW-12637/2020|EPI_ISL_570882|2020-0 ···················· ···················· ···················· ···················· ···················· ····················
K 2544 hCoV-19/USA/WA-UW-12652/2020|EPI_ISL_570883|2020-0 ···················· ···················· ···················· ···················· ···················· ····················
K 2545 hCoV-19/USA/WA-UW-12653/2020|EPI_ISL_570884|2020-0 ···················· ···················· ···················· ···················· ···················· ····················
K 2546 hCoV-19/USA/WA-UW-12658/2020|EPI_ISL_570885|2020-0 ···················· ···················· ···················· ···················· ···················· ····················
K 2547 hCoV-19/USA/WA-UW-12659/2020|EPI_ISL_570886|2020-0 ···················· ···················· ···················· ···················· ···················· ····················
K 2548 hCoV-19/USA/WA-UW-12660/2020|EPI_ISL_570887|2020-0 ···················· ···················· ···················· ···················· ···················· ····················
K 2549 hCoV-19/USA/WA-UW-12671/2020|EPI_ISL_570888|2020-0 ···················· ···················· ···················· ···················· ···················· ····················
K 2550 hCoV-19/USA/WA-UW-12685/2020|EPI_ISL_570889|2020-0 ···················· ···················· ···················· ···················· ···················· ····················
K 2551 hCoV-19/USA/WA-UW-12693/2020|EPI_ISL_570890|2020-0 ···················· ···················· ···················· ···················· ···················· ····················
K 2552 hCoV-19/USA/WA-UW-12709/2020|EPI_ISL_570891|2020-0 ···················· ···················· ···················· ···················· ···················· ····················
K 2553 hCoV-19/USA/WA-UW-12714/2020|EPI_ISL_570892|2020-0 ···················· ···················· ···················· ···················· ···················· ····················
K 2554 hCoV-19/USA/WA-UW-12716/2020|EPI_ISL_570893|2020-0 ···················· ···················· ···················· ···················· ···················· ····················
K 2555 hCoV-19/USA/WA-UW-12728/2020|EPI_ISL_570894|2020-0 ···················· ···················· ···················· ···················· ···················· ····················
K 2556 hCoV-19/USA/WA-UW-1273/2020|EPI_ISL_570895|2020-03 ···················· ···················· ···················· ···················· ···················· ····················
K 2557 hCoV-19/USA/WA-UW-12732/2020|EPI_ISL_570896|2020-0 ···················· ···················· ···················· ···················· ···················· ····················
K 2558 hCoV-19/USA/WA-UW-12733/2020|EPI_ISL_570897|2020-0 ···················· ···················· ···················· ···················· ···················· ····················
K 2559 hCoV-19/USA/WA-UW-12760/2020|EPI_ISL_570898|2020-0 ···················· ···················· ···················· ···················· ···················· ····················
K 2560 hCoV-19/USA/WA-UW-12761/2020|EPI_ISL_570899|2020-0 ···················· ···················· ···················· ···················· ···················· ····················
K 2562 hCoV-19/Gambia/GC19-2711/2020|EPI_ISL_561135|2020- ···················· ···················· ···················· ···················· ···················· ····················
K 2564 hCoV-19/Gambia/GC19-2731/2020|EPI_ISL_561137|2020- ···················· ···················· ···················· ···················· ···················· ····················
K 2565 hCoV-19/Gambia/GC192751/2020|EPI_ISL_561138|2020-0 ···················· ···················· ···················· ···················· ···················· ····················
K 2566 hCoV-19/Gambia/GC192756/2020|EPI_ISL_561139|2020-0 ···················· ···················· ···················· ···················· ···················· ····················
K 2567 hCoV-19/Gambia/GC192772/2020|EPI_ISL_561140|2020-0 ···················· ···················· ···················· ···················· ···················· ····················
K 2568 hCoV-19/Gambia/GC192778/2020|EPI_ISL_561141|2020-0 ···················· ···················· ···················· ···················· ···················· ····················
K 2569 hCoV-19/Gambia/GC192781/2020|EPI_ISL_561142|2020-0 ···················· ···················· ···················· ···················· ···················· ····················
K 2570 hCoV-19/Gambia/GC192782/2020|EPI_ISL_561143|2020-0 ···················· ···················· ···················· ···················· ···················· ····················
K 2571 hCoV-19/Gambia/GC192786/2020|EPI_ISL_561144|2020-0 ···················· ···················· ···················· ···················· ···················· ····················
K 2572 hCoV-19/Gambia/GC192789/2020|EPI_ISL_561145|2020-0 ···················· ···················· ···················· ···················· ···················· ····················
K 2573 hCoV-19/Gambia/GC192809/2020|EPI_ISL_561146|2020-0 ···················· ···················· ···················· ···················· ···················· ····················
K 2574 hCoV-19/Gambia/GC192812/2020|EPI_ISL_561148|2020-0 ···················· ···················· ···················· ···················· ···················· ····················
K 2575 hCoV-19/Gambia/GC19-2828/2020|EPI_ISL_561150|2020- ···················· ·············A······ ···················· ···················· ···················· ····················
K 2576 hCoV-19/Gambia/GC19-2842/2020|EPI_ISL_561151|2020- ···················· ···················· ···················· ···················· ···················· ····················
K 2577 hCoV-19/Gambia/GC19-2898/2020|EPI_ISL_561153|2020- ···················· ···················· ···················· ···················· ···················· ····················
K 2578 hCoV-19/Gambia/GC19-2904/2020|EPI_ISL_561154|2020- ···················· ···················· ···················· ···················· ···················· ····················
K 2579 hCoV-19/Gambia/GC19-2947/2020|EPI_ISL_561155|2020- ···················· ···················· ···················· ···················· ···················· ····················
K 2580 hCoV-19/Gambia/GC19-2973/2020|EPI_ISL_561156|2020- ···················· ···················· ···················· ···················· ···················· ····················
K 2581 hCoV-19/Gambia/GC19-2977/2020|EPI_ISL_561157|2020- ···················· ···················· ···················· ···················· ···················· ····················
K 2582 hCoV-19/Gambia/GC19-2985/2020|EPI_ISL_561158|2020- ···················· ···················· ···················· ···················· ···················· ····················
K 2583 hCoV-19/Gambia/GC19-2991/2020|EPI_ISL_561159|2020- ···················· ···················· ···················· ···················· ···················· ····················
K 2584 hCoV-19/Gambia/GC19-3064/2020|EPI_ISL_561160|2020- ···················· ···················· ···················· ···················· ···················· ····················
K 2585 hCoV-19/Gambia/GC19-3081/2020|EPI_ISL_561162|2020- ···················· ···················· ···················· ···················· ···················· ····················
K 2587 hCoV-19/Gambia/GC19-3099/2020|EPI_ISL_561164|2020- ···················· ···················· ···················· ···················· ···················· ····················
K 2588 hCoV-19/Gambia/GC19-3110/2020|EPI_ISL_561165|2020- ···················· ···················· ···················· ···················· ···················· ····················
K 2589 hCoV-19/Gambia/GC19-3112/2020|EPI_ISL_561166|2020- ···················· ···················· ···················· ···················· ···················· ····················
K 2590 hCoV-19/Gambia/GC19-3113/2020|EPI_ISL_561167|2020- ···················· ···················· ···················· ···················· ···················· ····················
K 2591 hCoV-19/Gambia/GC19-3114/2020|EPI_ISL_561168|2020- ···················· ···················· ···················· ···················· ···················· ····················
K 2593 hCoV-19/Gambia/GC19-3145/2020|EPI_ISL_561170|2020- ···················· ···················· ···················· ···················· ···················· ····················
K 2595 hCoV-19/Gambia/GC19-3399/2020|EPI_ISL_561172|2020- ···················· ···················· ···················· ···················· ···················· ····················
K 2596 hCoV-19/Gambia/GC19-3400/2020|EPI_ISL_561173|2020- ···················· ···················· ···················· ···················· ···················· ····················

| | | | | | | | | |
|---|---|---|---|---|---|---|---|---|
| K | 2597 | hCoV-19/Gambia/GC19-3407/2020\|EPI_ISL_561174\|2020- | ···················· | ···················· | ···················· | ···················· | ···················· | ···················· |
| K | 2598 | hCoV-19/Gambia/GC19-3418/2020\|EPI_ISL_561176\|2020- | ···················· | ···················· | ···················· | ···················· | ···················· | ···················· |
| K | 2599 | hCoV-19/Gambia/GC19-3423/2020\|EPI_ISL_561177\|2020- | ···················· | ···················· | ···················· | ···················· | ···················· | ···················· |
| K | 2600 | hCoV-19/Gambia/GC19-3429/2020\|EPI_ISL_561178\|2020- | ···················· | ···················· | ···················· | ···················· | ···················· | ···················· |
| K | 2602 | hCoV-19/Gambia/GC19-3438/2020\|EPI_ISL_561180\|2020- | ···················· | ···················· | ···················· | ···················· | ···················· | ···················· |
| K | 2603 | hCoV-19/Gambia/GC19-3445/2020\|EPI_ISL_561181\|2020- | ···················· | ···················· | ···················· | ···················· | ···················· | ···················· |
| K | 2604 | hCoV-19/Gambia/GC19-3447/2020\|EPI_ISL_561182\|2020- | ···················· | ···················· | ···················· | ···················· | ···················· | ···················· |
| K | 2605 | hCoV-19/Gambia/GC19-3497/2020\|EPI_ISL_561184\|2020- | ···················· | ···················· | ···················· | ···················· | ···················· | ···················· |
| K | 2606 | hCoV-19/Gambia/GC19-3501/2020\|EPI_ISL_561185\|2020- | ···················· | ···················· | ···················· | ···················· | ···················· | ···················· |
| K | 2608 | hCoV-19/Gambia/GC19-3505/2020\|EPI_ISL_561188\|2020- | ···················· | ···················· | ···················· | ···················· | ···················· | ···················· |
| K | 2609 | hCoV-19/Gambia/GC19-3514/2020\|EPI_ISL_561189\|2020- | ···················· | ···················· | ···················· | ···················· | ···················· | ···················· |
| K | 2610 | hCoV-19/Gambia/GC19-3528/2020\|EPI_ISL_561190\|2020- | ···················· | ···················· | ···················· | ···················· | ···················· | ···················· |
| K | 2611 | hCoV-19/Gambia/GC19-3529/2020\|EPI_ISL_561191\|2020- | ···················· | ···················· | ···················· | ···················· | ···················· | ···················· |
| K | 2612 | hCoV-19/Gambia/GC19-3533/2020\|EPI_ISL_561193\|2020- | ···················· | ···················· | ···················· | ···················· | ···················· | ···················· |
| K | 2613 | hCoV-19/Gambia/GC19-3535/2020\|EPI_ISL_561194\|2020- | ···················· | ···················· | ···················· | ···················· | ···················· | ···················· |
| K | 2614 | hCoV-19/Gambia/GC19-3550/2020\|EPI_ISL_561195\|2020- | ···················· | ···················· | ···················· | ···················· | ···················· | ···················· |
| K | 2615 | hCoV-19/Gambia/GC19-3565/2020\|EPI_ISL_561196\|2020- | ···················· | ···················· | ···················· | ···················· | ···················· | ···················· |
| K | 2616 | hCoV-19/Gambia/GC19-3613/2020\|EPI_ISL_561198\|2020- | ···················· | ···················· | ···················· | ···················· | ···················· | ···················· |
| K | 2617 | hCoV-19/Gambia/GC19-3629/2020\|EPI_ISL_561199\|2020- | ···················· | ···················· | ···················· | ···················· | ···················· | ···················· |
| K | 2618 | hCoV-19/Australia/VIC13388/2020\|EPI_ISL_563710\|202 | ···················· | ···················· | ···················· | ···················· | ···················· | ···················· |
| K | 2619 | hCoV-19/Australia/VIC13389/2020\|EPI_ISL_563711\|202 | ···················· | ···················· | ···················· | ···················· | ···················· | ···················· |
| K | 2620 | hCoV-19/Australia/VIC13392/2020\|EPI_ISL_563712\|202 | ···················· | ···················· | ···················· | ···················· | ···················· | ···················· |
| K | 2621 | hCoV-19/Australia/VIC13393/2020\|EPI_ISL_563713\|202 | ···················· | ···················· | ···················· | ···················· | ···················· | ···················· |
| K | 2622 | hCoV-19/Australia/VIC13397/2020\|EPI_ISL_563714\|202 | ···················· | ···················· | ···················· | ···················· | ···················· | ···················· |
| K | 2623 | hCoV-19/Australia/VIC13399/2020\|EPI_ISL_563715\|202 | ···················· | ···················· | ···················· | ···················· | ···················· | ···················· |
| K | 2624 | hCoV-19/Australia/VIC13400/2020\|EPI_ISL_563716\|202 | ···················· | ···················· | ···················· | ···················· | ···················· | ···················· |
| K | 2625 | hCoV-19/Australia/VIC13401/2020\|EPI_ISL_563717\|202 | ···················· | ···················· | ···················· | ···················· | ···················· | ···················· |
| K | 2626 | hCoV-19/Australia/VIC13403/2020\|EPI_ISL_563718\|202 | ···················· | ···················· | ···················· | ···················· | ···················· | ···················· |
| K | 2627 | hCoV-19/Australia/VIC13404/2020\|EPI_ISL_563719\|202 | ···················· | ···················· | ···················· | ···················· | ···················· | ···················· |
| K | 2628 | hCoV-19/Australia/VIC13406/2020\|EPI_ISL_563720\|202 | ···················· | ···················· | ···················· | ···················· | ···················· | ···················· |
| K | 2629 | hCoV-19/Australia/VIC13410/2020\|EPI_ISL_563721\|202 | ···················· | ···················· | ···················· | ···················· | ···················· | ···················· |
| K | 2630 | hCoV-19/Australia/VIC13415/2020\|EPI_ISL_563722\|202 | ···················· | ···················· | ··K················· | ···················· | ···················· | ···················· |
| K | 2631 | hCoV-19/Australia/VIC13419/2020\|EPI_ISL_563723\|202 | ···················· | ···················· | ···················· | ···················· | ···················· | ···················· |
| K | 2632 | hCoV-19/Australia/VIC13421/2020\|EPI_ISL_563724\|202 | ···················· | ···················· | ···················· | ···················· | ···················· | ···················· |
| K | 2633 | hCoV-19/Australia/VIC13423/2020\|EPI_ISL_563725\|202 | ···················· | ···················· | ···················· | ···················· | ···················· | ···················· |
| K | 2634 | hCoV-19/Australia/VIC13424/2020\|EPI_ISL_563726\|202 | ···················· | ···················· | ···················· | ···················· | ···················· | ···················· |
| K | 2635 | hCoV-19/Australia/VIC13425/2020\|EPI_ISL_563727\|202 | ···················· | ···················· | ···················· | ···················· | ···················· | ···················· |
| K | 2637 | hCoV-19/Australia/VIC13431/2020\|EPI_ISL_563729\|202 | ···················· | ···················· | ···················· | ···················· | ···················· | ···················· |
| K | 2638 | hCoV-19/Australia/VIC13538/2020\|EPI_ISL_563741\|202 | ···················· | ···················· | ···················· | ···················· | ···················· | ···················· |
| K | 2639 | hCoV-19/Australia/VIC13540/2020\|EPI_ISL_563742\|202 | ···················· | ···················· | ···················· | ···················· | ···················· | ···················· |
| K | 2640 | hCoV-19/Australia/VIC13542/2020\|EPI_ISL_563743\|202 | ···················· | ···················· | ···················· | ···················· | ···················· | ···················· |
| K | 2641 | hCoV-19/Australia/VIC13547/2020\|EPI_ISL_563745\|202 | ···················· | ···················· | ···················· | ···················· | ···················· | ···················· |
| K | 2642 | hCoV-19/Australia/VIC13548/2020\|EPI_ISL_563746\|202 | ···················· | ···················· | ···················· | ···················· | ···················· | ···················· |
| K | 2643 | hCoV-19/Australia/VIC13549/2020\|EPI_ISL_563747\|202 | ···················· | ···················· | ···················· | ···················· | ···················· | ···················· |
| K | 2644 | hCoV-19/Australia/VIC13551/2020\|EPI_ISL_563748\|202 | ···················· | ···················· | ···················· | ···················· | ···················· | ···················· |
| K | 2645 | hCoV-19/Australia/VIC13552/2020\|EPI_ISL_563749\|202 | ···················· | ···················· | ···················· | ···················· | ···················· | ···················· |
| K | 2646 | hCoV-19/Australia/VIC13652/2020\|EPI_ISL_563796\|202 | ···················· | ···················· | ···················· | ···················· | ···················· | ···················· |
| K | 2648 | hCoV-19/Australia/VIC13655/2020\|EPI_ISL_563798\|202 | ···················· | ···················· | ···················· | ···················· | ···················· | ···················· |
| K | 2649 | hCoV-19/Australia/VIC13656/2020\|EPI_ISL_563799\|202 | ···················· | ···················· | ···················· | ···················· | ···················· | ···················· |
| K | 2650 | hCoV-19/England/QEUH-9C8083/2020\|EPI_ISL_567200\|20 | ···················· | ···················· | ···················· | ···················· | ···················· | ···················· |
| K | 2651 | hCoV-19/England/QEUH-9C85B1/2020\|EPI_ISL_567201\|20 | ···················· | ···················· | ···················· | ···················· | ···················· | ···················· |
| K | 2652 | hCoV-19/England/QEUH-9C7B65/2020\|EPI_ISL_567203\|20 | ···················· | ···················· | ···················· | ···················· | ···················· | ···················· |
| K | 2653 | hCoV-19/England/QEUH-9C8A76/2020\|EPI_ISL_567204\|20 | ···················· | ···················· | ···················· | ···················· | ···················· | ···················· |
| K | 2654 | hCoV-19/England/QEUH-9C829C/2020\|EPI_ISL_567205\|20 | ···················· | ···················· | ···················· | ···················· | ···················· | ···················· |
| K | 2655 | hCoV-19/England/QEUH-9C80CF/2020\|EPI_ISL_567210\|20 | ···················· | ···················· | ···················· | ···················· | ···················· | ···················· |
| K | 2656 | hCoV-19/England/QEUH-9C8B91/2020\|EPI_ISL_567211\|20 | ···················· | ···················· | ···················· | ···················· | ···················· | ···················· |
| K | 2657 | hCoV-19/England/QEUH-9C7DBA/2020\|EPI_ISL_567212\|20 | ···················· | ···················· | ···················· | ···················· | ···················· | ···················· |
| K | 2658 | hCoV-19/England/QEUH-9C7BFC/2020\|EPI_ISL_567218\|20 | ···················· | ···················· | ···················· | ···················· | ···················· | ···················· |
| K | 2659 | hCoV-19/England/QEUH-9C8B19/2020\|EPI_ISL_567219\|20 | ···················· | ···················· | ···················· | ···················· | ···················· | ···················· |
| K | 2660 | hCoV-19/England/QEUH-9C89E2/2020\|EPI_ISL_567220\|20 | ···················· | ···················· | ···················· | ···················· | ···················· | ···················· |
| K | 2661 | hCoV-19/England/QEUH-9C9143/2020\|EPI_ISL_567222\|20 | ···················· | ···················· | ···················· | ···················· | ···················· | ···················· |
| K | 2662 | hCoV-19/England/QEUH-9C86EB/2020\|EPI_ISL_567223\|20 | ···················· | ···················· | ···················· | ···················· | ···················· | ···················· |
| K | 2663 | hCoV-19/England/QEUH-9C7D9C/2020\|EPI_ISL_567225\|20 | ···················· | ···················· | ···················· | ···················· | ·····T·············· | ···················· |
| K | 2664 | hCoV-19/England/QEUH-9C91E9/2020\|EPI_ISL_567226\|20 | ···················· | ···················· | ···················· | ···················· | ···················· | ···················· |
| K | 2665 | hCoV-19/England/QEUH-9C7EE4/2020\|EPI_ISL_567228\|20 | ···················· | ·A·················· | ···················· | ···················· | ···················· | ··A················· |
| K | 2666 | hCoV-19/England/QEUH-9C8DF5/2020\|EPI_ISL_567229\|20 | ···················· | ···················· | ···················· | ···················· | ···················· | ···················· |
| K | 2667 | hCoV-19/England/QEUH-9C7C35/2020\|EPI_ISL_567230\|20 | ···················· | ···················· | ···················· | ···················· | ···················· | ···················· |
| K | 2668 | hCoV-19/England/QEUH-9C83F3/2020\|EPI_ISL_567232\|20 | ···················· | ···················· | ···················· | ···················· | ···················· | ···················· |
| K | 2669 | hCoV-19/England/QEUH-9C8BFB/2020\|EPI_ISL_567233\|20 | ···················· | ···················· | ···················· | ···················· | ···················· | ···················· |
| K | 2670 | hCoV-19/England/QEUH-9C90BF/2020\|EPI_ISL_567235\|20 | ···················· | ···················· | ···················· | ···················· | ···················· | ···················· |
| K | 2671 | hCoV-19/England/QEUH-9C877F/2020\|EPI_ISL_567236\|20 | ···················· | ···················· | ···················· | ···················· | ···················· | ···················· |
| K | 2672 | hCoV-19/England/QEUH-9C836C/2020\|EPI_ISL_567237\|20 | ···················· | ···················· | ···················· | ···················· | ···················· | ···················· |
| K | 2673 | hCoV-19/England/QEUH-9C842D/2020\|EPI_ISL_567238\|20 | ···················· | ·A·················· | ···················· | ···················· | ···················· | ··A················· |
| K | 2674 | hCoV-19/Peru/LIM-UPCH-0130/2020\|EPI_ISL_568526\|202 | ···················· | ···················· | ···················· | ···················· | ···················· | ···················· |
| K | 2675 | hCoV-19/Peru/LIM-UPCH-0131/2020\|EPI_ISL_568527\|202 | ···················· | ···················· | ···················· | ···················· | ···················· | ···················· |
| K | 2676 | hCoV-19/Peru/LIM-UPCH-0132/2020\|EPI_ISL_568528\|202 | ···················· | ···················· | ···················· | ···················· | ···················· | ···················· |
| K | 2677 | hCoV-19/Peru/LIM-UPCH-0133/2020\|EPI_ISL_568529\|202 | ···················· | ···················· | ···················· | ···················· | ···················· | ···················· |
| K | 2678 | hCoV-19/Peru/LIM-UPCH-0134/2020\|EPI_ISL_568530\|202 | ···················· | ···················· | ···················· | ···················· | ···················· | ···················· |
| K | 2679 | hCoV-19/Peru/LIM-UPCH-0135/2020\|EPI_ISL_568531\|202 | ···················· | ···················· | ···················· | ···················· | ···················· | ···················· |
| K | 2680 | hCoV-19/Peru/LIM-UPCH-0136/2020\|EPI_ISL_568532\|202 | ···················· | ···················· | ···················· | ···················· | ···················· | ···················· |

```
K    2681 hCoV-19/Peru/LIM-UPCH-0137/2020|EPI_ISL_568533|202    ····················  ····················  ····················  ····················  ····················  ····················
K    2682 hCoV-19/Peru/LIM-UPCH-0138/2020|EPI_ISL_568534|202    ····················  ····················  ····················  ····················  ····················  ····················
K    2683 hCoV-19/Peru/LIM-UPCH-0141/2020|EPI_ISL_568535|202    ····················  ····················  ····················  ····················  ····················  ····················
K    2684 hCoV-19/Peru/LIM-UPCH-0142/2020|EPI_ISL_568536|202    ····················  ····················  ····················  ····················  ····················  ····················
K    2685 hCoV-19/Peru/LIM-UPCH-0146/2020|EPI_ISL_568540|202    ····················  ····················  ····················  ····················  ····················  ····················
K    2686 hCoV-19/Peru/LIM-UPCH-0147/2020|EPI_ISL_568541|202    ····················  ····················  ····················  ····················  ····················  ····················
K    2687 hCoV-19/Peru/LIM-UPCH-0148/2020|EPI_ISL_568542|202    ····················  ····················  ····················  ····················  ····················  ····················
K    2688 hCoV-19/Peru/LIM-UPCH-0149/2020|EPI_ISL_568543|202    ····················  ····················  ····················  ····················  ····················  ····················
K    2689 hCoV-19/Peru/LIM-UPCH-0155/2020|EPI_ISL_568548|202    ····················  ····················  ····················  ····················  ····················  ····················
K    2690 hCoV-19/Peru/LIM-UPCH-0156/2020|EPI_ISL_568549|202    ····················  ····················  ····················  ····················  ····················  ····················
K    2691 hCoV-19/Peru/LIM-UPCH-0157/2020|EPI_ISL_568550|202    ····················  ····················  ····················  ····················  ····················  ····················
K    2692 hCoV-19/Peru/LIM-UPCH-0158/2020|EPI_ISL_568551|202    ····················  ····················  ····················  ····················  ····················  ····················
K    2693 hCoV-19/Peru/LIM-UPCH-0159/2020|EPI_ISL_568552|202    ····················  ····················  ····················  ····················  ····················  ····················
K    2694 hCoV-19/Peru/LIM-UPCH-0160/2020|EPI_ISL_568553|202    ····················  ····················  ····················  ····················  ····················  ····················
K    2695 hCoV-19/Peru/LIM-UPCH-0161/2020|EPI_ISL_568554|202    ····················  ····················  ····················  ····················  ····················  ····················
K    2696 hCoV-19/Peru/LIM-UPCH-0162/2020|EPI_ISL_568555|202    ····················  ····················  ····················  ····················  ····················  ····················
K    2697 hCoV-19/Japan/NGY-NNH-001/2020|EPI_ISL_568556|2020    ····················  ····················  ····················  ····················  ····················  ····················
K    2698 hCoV-19/Japan/NGY-NNH-002/2020|EPI_ISL_568557|2020    ····················  ····················  ····················  ····················  ····················  ····················
K    2699 hCoV-19/Japan/NGY-NNH-003/2020|EPI_ISL_568558|2020    ····················  ····················  ····················  ····················  ····················  ····················
K    2700 hCoV-19/Japan/NGY-NNH-004/2020|EPI_ISL_568559|2020    ····················  ····················  ····················  ····················  ····················  ····················
K    2701 hCoV-19/Japan/NGY-NNH-005/2020|EPI_ISL_568560|2020    ····················  ····················  ····················  ····················  ····················  ····················
K    2702 hCoV-19/Japan/NGY-NNH-006/2020|EPI_ISL_568561|2020    ····················  ····················  ····················  ····················  ····················  ····················
K    2703 hCoV-19/Japan/NGY-NNH-007/2020|EPI_ISL_568562|2020    ····················  ····················  ····················  ····················  ····················  ····················
K    2704 hCoV-19/Japan/NGY-NNH-008/2020|EPI_ISL_568563|2020    ····················  ····················  ····················  ····················  ····················  ····················
K    2705 hCoV-19/Japan/NGY-NNH-009/2020|EPI_ISL_568564|2020    ····················  ····················  ····················  ····················  ····················  ····················
K    2706 hCoV-19/Japan/NGY-NNH-010/2020|EPI_ISL_568565|2020    ····················  ····················  ····················  ····················  ····················  ····················
K    2707 hCoV-19/Japan/NGY-NNH-011/2020|EPI_ISL_568566|2020    ····················  ····················  ····················  ····················  ····················  ····················
K    2708 hCoV-19/Japan/NGY-NNH-012/2020|EPI_ISL_568567|2020    ····················  ····················  ····················  ····················  ····················  ···············A····
K    2709 hCoV-19/Japan/NGY-NNH-013/2020|EPI_ISL_568568|2020    ····················  ····················  ····················  ····················  ····················  ····················
K    2710 hCoV-19/Japan/NGY-NNH-014/2020|EPI_ISL_568569|2020    ····················  ····················  ····················  ····················  ····················  ····················
K    2711 hCoV-19/USA/FL-BPHL-1502/2020|EPI_ISL_568600|2020-    ····················  ····················  ····················  ····················  ····················  ····················
K    2713 hCoV-19/USA/FL-BPHL-1504/2020|EPI_ISL_568602|2020-    ····················  ····················  ····················  ····················  ····················  ····················
K    2714 hCoV-19/Russia/OMS-ORINFI-4895S/2020|EPI_ISL_56985    ····················  ····················  ····················  ····················  ····················  ····················
K    2715 hCoV-19/Australia/NSW2755/2020|EPI_ISL_569858|2020    ····················  ····················  ····················  ····················  ····················  ····················
K    2716 hCoV-19/India/WB-IICB-001/2020|EPI_ISL_569859|2020    ····················  ·A··················  ····················  ····················  ····················  ···A················
K    2717 hCoV-19/India/WB-IICB-002/2020|EPI_ISL_569860|2020    ····················  ·A··················  ····················  ····················  ····················  ····················
K    2718 hCoV-19/India/WB-IICB-003/2020|EPI_ISL_569861|2020    ····················  ····················  ····················  ····················  ····················  ····················
K    2719 hCoV-19/India/WB-IICB-004/2020|EPI_ISL_569862|2020    ····················  ·A··················  ····················  ····················  ····················  ····················
K    2720 hCoV-19/India/WB-IICB-005/2020|EPI_ISL_569863|2020    ····················  ····················  ····················  ····················  ····················  ····················
K    2721 hCoV-19/Italy/PIE-SLL-NZ145/2020|EPI_ISL_569865|20    ····················  ····················  ····················  ····················  ····················  ····················
K    2722 hCoV-19/Italy/PIE-SLL-MS45/2020|EPI_ISL_569866|202    ····················  ····················  ····················  ····················  ····················  ····················
K    2723 hCoV-19/Italy/PIE-SLL-MS46/2020|EPI_ISL_569867|202    ····················  ····················  ····················  ····················  ····················  ····················
K    2724 hCoV-19/Italy/PIE-SLL-MS54/2020|EPI_ISL_569870|202    ····················  ····················  ····················  ····················  ····················  ····················
K    2725 hCoV-19/Italy/PIE-SLL-MS55/2020|EPI_ISL_569871|202    ····················  ····················  ····················  ····················  ····················  ····················
K    2726 hCoV-19/Italy/PIE-SLL-MS56/2020|EPI_ISL_569872|202    ····················  ····················  ····················  ····················  ····················  ····················
K    2727 hCoV-19/Italy/PIE-SLL-MS57/2020|EPI_ISL_569873|202    ····················  ····················  ····················  ····················  ····················  ····················
K    2728 hCoV-19/Italy/PIE-SLL-MS58/2020|EPI_ISL_569874|202    ····················  ····················  ····················  ····················  ····················  ····················
K    2729 hCoV-19/Italy/PIE-SLL-NZ138/2020|EPI_ISL_569879|20    ····················  ····················  ····················  ····················  ····················  ····················
K    2730 hCoV-19/Italy/PIE-SLL-NZ140/2020|EPI_ISL_569880|20    ····················  ····················  ····················  ····················  ····················  ····················
K    2731 hCoV-19/Italy/PIE-SLL-NZ141/2020|EPI_ISL_569881|20    ····················  ····················  ····················  ····················  ····················  ····················
K    2732 hCoV-19/Italy/PIE-SLL-NZ142/2020|EPI_ISL_569882|20    ····················  ····················  ····················  ····················  ····················  ····················
K    2733 hCoV-19/Italy/PIE-SLL-NZ144/2020|EPI_ISL_569883|20    ····················  ····················  ····················  ····················  ····················  ····················
K    2734 hCoV-19/Italy/PIE-SLL-NZ146/2020|EPI_ISL_569884|20    ····················  ····················  ····················  ····················  ····················  ····················
K    2735 hCoV-19/Italy/PIE-SLL-NZ147/2020|EPI_ISL_569885|20    ····················  ····················  ····················  ····················  ····················  ····················
K    2736 hCoV-19/Italy/PIE-SLL-NZ148/2020|EPI_ISL_569886|20    ····················  ····················  ····················  ····················  ····················  ····················
K    2737 hCoV-19/USA/CA-IGI-0202/2020|EPI_ISL_569887|2020-0    ····················  ····················  ····················  ····················  ····················  ····················
K    2738 hCoV-19/USA/CA-IGI-0203/2020|EPI_ISL_569888|2020-0    ····················  ····················  ····················  ····················  ····················  ····················
K    2739 hCoV-19/USA/CA-IGI-0205/2020|EPI_ISL_569889|2020-0    ····················  ····················  ····················  ····················  ····················  ····················
K    2740 hCoV-19/USA/CA-IGI-0209/2020|EPI_ISL_569890|2020-0    ····················  ····················  ····················  ····················  ····················  ····················
K    2741 hCoV-19/USA/CA-IGI-0210/2020|EPI_ISL_569891|2020-0    ····················  ····················  ····················  ····················  ····················  ····················
K    2742 hCoV-19/USA/CA-IGI-0211/2020|EPI_ISL_569892|2020-0    ····················  ····················  ····················  ····················  ····················  ····················
K    2743 hCoV-19/USA/CA-IGI-0214/2020|EPI_ISL_569893|2020-0    ····················  ····················  ····················  ····················  ····················  ····················
K    2744 hCoV-19/USA/CA-IGI-0216/2020|EPI_ISL_569894|2020-0    ····················  ····················  ····················  ····················  ····················  ····················
K    2745 hCoV-19/USA/CA-IGI-0218/2020|EPI_ISL_569895|2020-0    ····················  ····················  ····················  ····················  ····················  ····················
K    2746 hCoV-19/USA/CA-IGI-0220/2020|EPI_ISL_569896|2020-0    ····················  ····················  ····················  ····················  ····················  ····················
K    2747 hCoV-19/USA/CA-IGI-0224/2020|EPI_ISL_569898|2020-0    ····················  ·A··················  ····················  ····················  ····················  ····················
K    2748 hCoV-19/USA/CA-IGI-0225/2020|EPI_ISL_569899|2020-0    ····················  ····················  ····················  ····················  ····················  ····················
K    2749 hCoV-19/USA/CA-IGI-0228/2020|EPI_ISL_569900|2020-0    ····················  ····················  ····················  ····················  ····················  ····················
K    2750 hCoV-19/USA/CA-IGI-0229/2020|EPI_ISL_569901|2020-0    ····················  ····················  ····················  ····················  ····················  ····················
K    2751 hCoV-19/USA/CA-IGI-0230/2020|EPI_ISL_569902|2020-0    ····················  ····················  ····················  ····················  ····················  ····················
K    2752 hCoV-19/USA/CA-IGI-0235/2020|EPI_ISL_569904|2020-0    ····················  ·A··················  ····················  ····················  ····················  ····················
K    2753 hCoV-19/USA/CA-IGI-0236/2020|EPI_ISL_569905|2020-0    ····················  ····················  ····················  ····················  ····················  ····················
K    2754 hCoV-19/USA/CA-IGI-0237/2020|EPI_ISL_569906|2020-0    ····················  ·A··················  ····················  ····················  ····················  ····················
K    2755 hCoV-19/USA/CA-IGI-0240/2020|EPI_ISL_569907|2020-0    ····················  ····················  ····················  ····················  ····················  ····················
K    2756 hCoV-19/USA/CA-IGI-0245/2020|EPI_ISL_569909|2020-0    ····················  ····················  ····················  ····················  ····················  ····················
K    2757 hCoV-19/USA/CA-IGI-0248/2020|EPI_ISL_569911|2020-0    ····················  ····················  ····················  ····················  ····················  ····················
K    2758 hCoV-19/USA/CA-IGI-0250/2020|EPI_ISL_569913|2020-0    ····················  ····················  ····················  ····················  ····················  ····················
K    2759 hCoV-19/USA/CA-IGI-0253/2020|EPI_ISL_569915|2020-0    ····················  ····················  ····················  ····················  ····················  ····················
K    2760 hCoV-19/USA/CA-IGI-0204/2020|EPI_ISL_569932|2020-0    ····················  ····················  ····················  ····················  ····················  ····················
K    2761 hCoV-19/USA/CA-IGI-0206/2020|EPI_ISL_569933|2020-0    ····················  ····················  ····················  ····················  ····················  ····················
```

| | | | | | | | | |
|---|---|---|---|---|---|---|---|---|
| K | 2762 | hCoV-19/USA/WA-UW-21759/2020\|EPI_ISL_570840\|2020-0 | ···················· | ···················· | ···················· | ···················· | ···················· | ···················· |
| K | 2763 | hCoV-19/USA/WA-UW-12796/2020\|EPI_ISL_570841\|2020-0 | ···················· | ···················· | ···················· | ···················· | ···················· | ···················· |
| K | 2764 | hCoV-19/USA/OR-UW-12762/2020\|EPI_ISL_570842\|2020-0 | ···················· | ···················· | ···················· | ···················· | ···················· | ···················· |
| K | 2765 | hCoV-19/USA/WA-UW-21409/2020\|EPI_ISL_570843\|2020-0 | ···················· | ·A·················· | ···················· | ···················· | ···················· | ···················· |
| K | 2766 | hCoV-19/USA/WA-UW-13047/2020\|EPI_ISL_570844\|2020-0 | ···················· | ···················· | ···················· | ···················· | ···················· | ···················· |
| K | 2767 | hCoV-19/USA/WA-UW-21721/2020\|EPI_ISL_570846\|2020-0 | ···················· | ·A·················· | ···················· | ···················· | ···················· | ···················· |
| K | 2768 | hCoV-19/USA/WA-UW-12899/2020\|EPI_ISL_570847\|2020-0 | ···················· | ···················· | ···················· | ···················· | ···················· | ···················· |
| K | 2769 | hCoV-19/USA/ID-UW-21755/2020\|EPI_ISL_570848\|2020-0 | ···················· | ···················· | ···················· | ···················· | ···················· | ···················· |
| K | 2770 | hCoV-19/USA/OR-UW-12646/2020\|EPI_ISL_570849\|2020-0 | ···················· | ···················· | ···················· | ···················· | ···················· | ···················· |
| K | 2771 | hCoV-19/USA/WA-UW-13031/2020\|EPI_ISL_570850\|2020-0 | ···················· | ···················· | ···················· | ···················· | ···················· | ···················· |
| K | 2772 | hCoV-19/USA/WA-UW-13081/2020\|EPI_ISL_570851\|2020-0 | ···················· | ···················· | ···················· | ···················· | ···················· | ···················· |
| K | 2773 | hCoV-19/USA/WA-UW-19260/2020\|EPI_ISL_570852\|2020-0 | ···················· | ·A·················· | ···················· | ···················· | ···················· | ···················· |
| K | 2774 | hCoV-19/USA/WA-UW-19262/2020\|EPI_ISL_570853\|2020-0 | ···················· | ···················· | ···················· | ···················· | ···················· | ···················· |
| K | 2775 | hCoV-19/USA/WA-UW-19376/2020\|EPI_ISL_570854\|2020-0 | ···················· | ···················· | ···················· | ···················· | ···················· | ···················· |
| K | 2776 | hCoV-19/USA/WA-UW-12641/2020\|EPI_ISL_570859\|2020-0 | ···················· | ···················· | ···················· | ···················· | ···················· | ···················· |
| K | 2777 | hCoV-19/USA/WA-UW-13039/2020\|EPI_ISL_570860\|2020-0 | ···················· | ···················· | ···················· | ···················· | ···················· | ···················· |
| K | 2778 | hCoV-19/USA/WA-UW-21728/2020\|EPI_ISL_570861\|2020-0 | ···················· | ···················· | ···················· | ···················· | ···················· | ···················· |
| K | 2779 | hCoV-19/USA/OR-UW-12645/2020\|EPI_ISL_570862\|2020-0 | ···················· | ···················· | ···················· | ···················· | ···················· | ···················· |
| K | 2780 | hCoV-19/USA/WA-UW-12557/2020\|EPI_ISL_570863\|2020-0 | ···················· | ·A·················· | ···················· | ···················· | ···················· | ···················· |
| K | 2781 | hCoV-19/USA/WA-UW-12570/2020\|EPI_ISL_570864\|2020-0 | ···················· | ···················· | ···················· | ···················· | ···················· | ···················· |
| K | 2782 | hCoV-19/USA/WA-UW-12575/2020\|EPI_ISL_570865\|2020-0 | ···················· | ···················· | ···················· | ···················· | ···················· | ···················· |
| K | 2783 | hCoV-19/USA/WA-UW-12576/2020\|EPI_ISL_570866\|2020-0 | ···················· | ···················· | ···················· | ···················· | ···················· | ···················· |
| K | 2784 | hCoV-19/USA/WA-UW-12578/2020\|EPI_ISL_570867\|2020-0 | ···················· | ···················· | ···················· | ···················· | ···················· | ···················· |
| K | 2785 | hCoV-19/USA/WA-UW-12582/2020\|EPI_ISL_570868\|2020-0 | ···················· | ···················· | ···················· | ···················· | ···················· | ···················· |
| K | 2786 | hCoV-19/USA/WA-UW-12593/2020\|EPI_ISL_570869\|2020-0 | ···················· | ···················· | ···················· | ···················· | ···················· | ···················· |
| K | 2787 | hCoV-19/USA/WA-UW-12594/2020\|EPI_ISL_570870\|2020-0 | ···················· | ·A·················· | ···················· | ···················· | ···················· | ···················· |
| K | 2788 | hCoV-19/USA/WA-UW-12600/2020\|EPI_ISL_570871\|2020-0 | ···················· | ···················· | ···················· | ···················· | ···················· | ···················· |
| K | 2789 | hCoV-19/USA/WA-UW-12602/2020\|EPI_ISL_570872\|2020-0 | ···················· | ·A·················· | ···················· | ···················· | ···················· | ···················· |
| K | 2790 | hCoV-19/USA/WA-UW-12604/2020\|EPI_ISL_570873\|2020-0 | ···················· | ···················· | ···················· | ···················· | ···················· | ···················· |
| K | 2791 | hCoV-19/USA/WA-UW-12611/2020\|EPI_ISL_570874\|2020-0 | ···················· | ···················· | ···················· | ···················· | ···················· | ···················· |
| K | 2792 | hCoV-19/USA/WA-UW-12613/2020\|EPI_ISL_570875\|2020-0 | ···················· | ···················· | ···················· | ···················· | ···················· | ···················· |
| K | 2793 | hCoV-19/USA/WA-UW-12621/2020\|EPI_ISL_570876\|2020-0 | ···················· | ···················· | ···················· | ···················· | ···················· | ···················· |
| K | 2794 | hCoV-19/USA/WA-UW-12624/2020\|EPI_ISL_570877\|2020-0 | ···················· | ·A·················· | ···················· | ···················· | ···················· | ···················· |
| K | 2795 | hCoV-19/USA/WA-UW-12625/2020\|EPI_ISL_570878\|2020-0 | ···················· | ···················· | ···················· | ···················· | ···················· | ···················· |
| K | 2796 | hCoV-19/USA/WA-UW-12626/2020\|EPI_ISL_570879\|2020-0 | ···················· | ···················· | ···················· | ···················· | ···················· | ···················· |
| K | 2797 | hCoV-19/USA/WA-UW-12763/2020\|EPI_ISL_570900\|2020-0 | ···················· | ···················· | ···················· | ···················· | ···················· | ···················· |
| K | 2798 | hCoV-19/USA/WA-UW-1278/2020\|EPI_ISL_570901\|2020-03 | ···················· | ···················· | ···················· | ···················· | ···················· | ···················· |
| K | 2799 | hCoV-19/USA/WA-UW-12785/2020\|EPI_ISL_570902\|2020-0 | ···················· | ·A·················· | ···················· | ···················· | ···················· | ···················· |
| K | 2800 | hCoV-19/USA/WA-UW-12797/2020\|EPI_ISL_570903\|2020-0 | ···················· | ···················· | ···················· | ···················· | ···················· | ···················· |
| K | 2801 | hCoV-19/USA/WA-UW-12801/2020\|EPI_ISL_570904\|2020-0 | ···················· | ···················· | ···················· | ···················· | ···················· | ···················· |
| K | 2802 | hCoV-19/USA/WA-UW-12822/2020\|EPI_ISL_570905\|2020-0 | ···················· | ···················· | ···················· | ···················· | ···················· | ···················· |
| K | 2803 | hCoV-19/USA/WA-UW-12828/2020\|EPI_ISL_570906\|2020-0 | ···················· | ···················· | ···················· | ···················· | ···················· | ···················· |
| K | 2804 | hCoV-19/USA/WA-UW-12830/2020\|EPI_ISL_570907\|2020-0 | ···················· | ···················· | ···················· | ···················· | ···················· | ···················· |
| K | 2805 | hCoV-19/USA/WA-UW-12832/2020\|EPI_ISL_570908\|2020-0 | ···················· | ·A·················· | ···················· | ···················· | ···················· | ···················· |
| K | 2806 | hCoV-19/USA/WA-UW-12835/2020\|EPI_ISL_570909\|2020-0 | ···················· | ···················· | ···················· | ···················· | ···················· | ···················· |
| K | 2807 | hCoV-19/USA/WA-UW-12839/2020\|EPI_ISL_570910\|2020-0 | ···················· | ·A·················· | ···················· | ···················· | ···················· | ···················· |
| K | 2808 | hCoV-19/USA/WA-UW-12846/2020\|EPI_ISL_570911\|2020-0 | ···················· | ···················· | ···················· | ···················· | ···················· | ···················· |
| K | 2809 | hCoV-19/USA/WA-UW-1285/2020\|EPI_ISL_570912\|2020-03 | ···················· | ···················· | ···················· | ···················· | ···················· | ···················· |
| K | 2810 | hCoV-19/USA/WA-UW-12853/2020\|EPI_ISL_570913\|2020-0 | ···················· | ···················· | ···················· | ···················· | ···················· | ···················· |
| K | 2811 | hCoV-19/USA/WA-UW-12858/2020\|EPI_ISL_570914\|2020-0 | ···················· | ···················· | ···················· | ···················· | ···················· | ···················· |
| K | 2812 | hCoV-19/USA/WA-UW-12861/2020\|EPI_ISL_570915\|2020-0 | ···················· | ···················· | ···················· | ···················· | ···················· | ···················· |
| K | 2813 | hCoV-19/USA/WA-UW-12863/2020\|EPI_ISL_570916\|2020-0 | ···················· | ·A·················· | ···················· | ···················· | ···················· | ···················· |
| K | 2814 | hCoV-19/USA/WA-UW-12875/2020\|EPI_ISL_570917\|2020-0 | ···················· | ·A·················· | ···················· | ···················· | ···················· | ···················· |
| K | 2815 | hCoV-19/USA/WA-UW-12876/2020\|EPI_ISL_570918\|2020-0 | ···················· | ·A·················· | ···················· | ···················· | ···················· | ···················· |
| K | 2816 | hCoV-19/USA/WA-UW-12878/2020\|EPI_ISL_570919\|2020-0 | ···················· | ···················· | ···················· | ···················· | ···················· | ···················· |
| K | 2817 | hCoV-19/Gambia/GC19-3846/2020\|EPI_ISL_561100\|2020- | ···················· | ···················· | ···················· | ···················· | ···················· | ···················· |
| K | 2818 | hCoV-19/Gambia/GC19-3854/2020\|EPI_ISL_561102\|2020- | ···················· | ···················· | ···················· | ···················· | ···················· | ···················· |
| K | 2819 | hCoV-19/Gambia/GC19-2427/2020\|EPI_ISL_561105\|2020- | ···················· | ···················· | ···················· | ···················· | ···················· | ···················· |
| K | 2820 | hCoV-19/Gambia/GC19-2450/2020\|EPI_ISL_561107\|2020- | ···················· | ···················· | ···················· | ···················· | ···················· | ···················· |
| K | 2821 | hCoV-19/Gambia/GC19-2459/2020\|EPI_ISL_561108\|2020- | ···················· | ···················· | ···················· | ···················· | ···················· | ···················· |
| K | 2822 | hCoV-19/Gambia/GC19-2473/2020\|EPI_ISL_561110\|2020- | ···················· | ···················· | ···················· | ···················· | ···················· | ···················· |
| K | 2823 | hCoV-19/Gambia/GC19-2498/2020\|EPI_ISL_561111\|2020- | ···················· | ···················· | ···················· | ···················· | ···················· | ···················· |
| K | 2824 | hCoV-19/Gambia/GC19-2501/2020\|EPI_ISL_561112\|2020- | ···················· | ···················· | ···················· | ···················· | ···················· | ···················· |
| K | 2825 | hCoV-19/Gambia/GC19-2525/2020\|EPI_ISL_561113\|2020- | ···················· | ···················· | ···················· | ···················· | ···················· | ···················· |
| K | 2826 | hCoV-19/Gambia/GC19-2526/2020\|EPI_ISL_561114\|2020- | ···················· | ···················· | ···················· | ···················· | ···················· | ···················· |
| K | 2827 | hCoV-19/Gambia/GC192573/2020\|EPI_ISL_561115\|2020-0 | ···················· | ···················· | ···················· | ···················· | ···················· | ···················· |
| K | 2828 | hCoV-19/Gambia/GC192574/2020\|EPI_ISL_561116\|2020-0 | ···················· | ···················· | ···················· | ···················· | ···················· | ···················· |
| K | 2829 | hCoV-19/Gambia/GC192576/2020\|EPI_ISL_561117\|2020-0 | ···················· | ···················· | ···················· | ···················· | ···················· | ···················· |
| K | 2830 | hCoV-19/Gambia/GC192579/2020\|EPI_ISL_561118\|2020-0 | ···················· | ···················· | ···················· | ···················· | ···················· | ···················· |
| K | 2831 | hCoV-19/Gambia/GC192583/2020\|EPI_ISL_561119\|2020-0 | ···················· | ···················· | ···················· | ···················· | ···················· | ···················· |
| K | 2832 | hCoV-19/Gambia/GC192602/2020\|EPI_ISL_561120\|2020-0 | ···················· | ···················· | ···················· | ···················· | ···················· | ···················· |
| K | 2833 | hCoV-19/Gambia/GC192610/2020\|EPI_ISL_561121\|2020-0 | ···················· | ···················· | ···················· | ···················· | ···················· | ···················· |
| K | 2834 | hCoV-19/Gambia/GC19-2628/2020\|EPI_ISL_561122\|2020- | ···················· | ···················· | ···················· | ···················· | ···················· | ···················· |
| K | 2835 | hCoV-19/Gambia/GC19-2629/2020\|EPI_ISL_561123\|2020- | ···················· | ···················· | ···················· | ···················· | ···················· | ···················· |
| K | 2837 | hCoV-19/Gambia/GC19-2637/2020\|EPI_ISL_561125\|2020- | ···················· | ···················· | ···················· | ···················· | ···················· | ···················· |
| K | 2838 | hCoV-19/Gambia/GC19-2641/2020\|EPI_ISL_561127\|2020- | ···················· | ···················· | ···················· | ···················· | ···················· | ···················· |
| K | 2839 | hCoV-19/Gambia/GC19-2653/2020\|EPI_ISL_561129\|2020- | ···················· | ···················· | ···················· | ···················· | ···················· | ···················· |
| K | 2840 | hCoV-19/Gambia/GC19-2654/2020\|EPI_ISL_561130\|2020- | ···················· | ···················· | ···················· | ···················· | ···················· | ···················· |
| K | 2841 | hCoV-19/Gambia/GC19-2660/2020\|EPI_ISL_561131\|2020- | ···················· | ···················· | ···················· | ···················· | ···················· | ···················· |
| K | 2842 | hCoV-19/Gambia/GC19-2662/2020\|EPI_ISL_561132\|2020- | ···················· | ···················· | ···················· | ···················· | ···················· | ···················· |

| | | | | | | | | |
|---|---|---|---|---|---|---|---|---|
| K | 2843 | hCoV-19/Gambia/GC19-2663/2020\|EPI_ISL_561133\|2020- | .................... | .................... | ..................... | .................... | .................... | .................... |
| K | 2844 | hCoV-19/Australia/VIC13432/2020\|EPI_ISL_563730\|202 | .................... | .................... | ..................... | .................... | .................... | .................... |
| K | 2845 | hCoV-19/Australia/VIC13433/2020\|EPI_ISL_563731\|202 | .................... | .................... | ..................... | .................... | .................... | .................... |
| K | 2846 | hCoV-19/Australia/VIC13436/2020\|EPI_ISL_563732\|202 | .................... | .................... | ..................... | .................... | .................... | .................... |
| K | 2847 | hCoV-19/Australia/VIC13437/2020\|EPI_ISL_563733\|202 | .................... | .................... | ..................... | .................... | .................... | .................... |
| K | 2848 | hCoV-19/Australia/VIC13439/2020\|EPI_ISL_563734\|202 | .................... | .................... | ..................... | .................... | .................... | .................... |
| K | 2849 | hCoV-19/Australia/VIC13446/2020\|EPI_ISL_563735\|202 | .................... | .................... | ..................... | .................... | .................... | .................... |
| K | 2850 | hCoV-19/Australia/VIC13448/2020\|EPI_ISL_563736\|202 | .................... | .................... | ..................... | .................... | .................... | .................... |
| K | 2851 | hCoV-19/Australia/VIC13461/2020\|EPI_ISL_563737\|202 | .................... | .................... | ..................... | .................... | .................... | .................... |
| K | 2853 | hCoV-19/Australia/VIC13535/2020\|EPI_ISL_563739\|202 | .................... | .................... | ..................... | .................... | .................... | .................... |
| K | 2854 | hCoV-19/Australia/VIC13536/2020\|EPI_ISL_563740\|202 | .................... | .................... | ..................... | .................... | .................... | .................... |
| K | 2855 | hCoV-19/Australia/VIC13545/2020\|EPI_ISL_563744\|202 | .................... | .................... | ..................... | .................... | .................... | .................... |
| K | 2856 | hCoV-19/Australia/VIC13553/2020\|EPI_ISL_563750\|202 | .................... | .................... | ..................... | .................... | .................... | .................... |
| K | 2857 | hCoV-19/Australia/VIC13554/2020\|EPI_ISL_563751\|202 | .................... | .................... | ..................... | .................... | .................... | .................... |
| K | 2858 | hCoV-19/Australia/VIC13556/2020\|EPI_ISL_563752\|202 | .................... | .................... | ..................... | .................... | .................... | .................... |
| K | 2859 | hCoV-19/Australia/VIC13558/2020\|EPI_ISL_563753\|202 | .................... | .................... | ..................... | .................... | .................... | .................... |
| K | 2860 | hCoV-19/Australia/VIC13559/2020\|EPI_ISL_563754\|202 | .................... | .................... | ..................... | .................... | .................... | .................... |
| K | 2861 | hCoV-19/Australia/VIC13563/2020\|EPI_ISL_563755\|202 | .................... | .................... | ..................... | .................... | .................... | .................... |
| K | 2862 | hCoV-19/Australia/VIC13565/2020\|EPI_ISL_563756\|202 | .................... | .................... | ..................... | .................... | .................... | .................... |
| K | 2863 | hCoV-19/Australia/VIC13566/2020\|EPI_ISL_563757\|202 | .................... | .................... | ..................... | .................... | .................... | .................... |
| K | 2864 | hCoV-19/Australia/VIC13569/2020\|EPI_ISL_563758\|202 | .................... | .................... | ..................... | .................... | .................... | .................... |
| K | 2865 | hCoV-19/Australia/VIC13570/2020\|EPI_ISL_563759\|202 | .................... | .................... | ..................... | .................... | .................... | .................... |
| K | 2866 | hCoV-19/Australia/VIC13572/2020\|EPI_ISL_563760\|202 | .................... | .................... | ..................... | .................... | .................... | .................... |
| K | 2867 | hCoV-19/Australia/VIC13573/2020\|EPI_ISL_563761\|202 | .................... | .................... | ..................... | .................... | .................... | .................... |
| K | 2868 | hCoV-19/Australia/VIC13575/2020\|EPI_ISL_563762\|202 | .................... | .................... | ..................... | .................... | .................... | .................... |
| K | 2869 | hCoV-19/Australia/VIC13576/2020\|EPI_ISL_563763\|202 | .................... | .................... | ...........T......... | .................... | .................... | .................... |
| K | 2870 | hCoV-19/Australia/VIC13581/2020\|EPI_ISL_563764\|202 | .................... | .................... | ..................... | .................... | .................... | .................... |
| K | 2871 | hCoV-19/Australia/VIC13582/2020\|EPI_ISL_563765\|202 | .................... | .................... | ..................... | .................... | .................... | .................... |
| K | 2872 | hCoV-19/Australia/VIC13583/2020\|EPI_ISL_563766\|202 | .................... | .................... | ..................... | .................... | .................... | .................... |
| K | 2873 | hCoV-19/Australia/VIC13585/2020\|EPI_ISL_563767\|202 | .................... | .................... | ..................... | .................... | .................... | .................... |
| K | 2874 | hCoV-19/Australia/VIC13586/2020\|EPI_ISL_563768\|202 | .................... | .................... | ..................... | .................... | .................... | .................... |
| K | 2875 | hCoV-19/Australia/VIC13587/2020\|EPI_ISL_563769\|202 | .................... | .................... | ..................... | .................... | .................... | .................... |
| K | 2876 | hCoV-19/Australia/VIC13591/2020\|EPI_ISL_563770\|202 | .................... | .................... | ..................... | .................... | .................... | .................... |
| K | 2877 | hCoV-19/Australia/VIC13593/2020\|EPI_ISL_563771\|202 | .................... | .................... | ..................... | .................... | .................... | .................... |
| K | 2878 | hCoV-19/Australia/VIC13594/2020\|EPI_ISL_563772\|202 | .................... | .................... | ..................... | .................... | .................... | .................... |
| K | 2879 | hCoV-19/Australia/VIC13595/2020\|EPI_ISL_563773\|202 | .................... | .................... | ..................... | .................... | .................... | .................... |
| K | 2880 | hCoV-19/Australia/VIC13596/2020\|EPI_ISL_563774\|202 | .................... | .................... | ..................... | .................... | .................... | .................... |
| K | 2881 | hCoV-19/Australia/VIC13599/2020\|EPI_ISL_563775\|202 | .................... | .................... | ..................... | .................... | .................... | .................... |
| K | 2882 | hCoV-19/Australia/VIC13600/2020\|EPI_ISL_563776\|202 | .................... | .................... | ..................... | .................... | .................... | .................... |
| K | 2883 | hCoV-19/Australia/VIC13601/2020\|EPI_ISL_563777\|202 | .................... | .................... | ..................... | .................... | .................... | .................... |
| K | 2884 | hCoV-19/Australia/VIC13602/2020\|EPI_ISL_563778\|202 | .................... | .................... | ..................... | .................... | .................... | .................... |
| K | 2885 | hCoV-19/Australia/VIC13603/2020\|EPI_ISL_563779\|202 | .................... | .................... | ..................... | .................... | .................... | .................... |
| K | 2886 | hCoV-19/Australia/VIC13604/2020\|EPI_ISL_563780\|202 | .................... | .................... | ..................... | .................... | .................... | .................... |
| K | 2887 | hCoV-19/Australia/VIC13606/2020\|EPI_ISL_563781\|202 | .................... | .................... | ..................... | .................... | .................... | .................... |
| K | 2888 | hCoV-19/Australia/VIC13607/2020\|EPI_ISL_563782\|202 | .................... | .................... | ..................... | .................... | .................... | .................... |
| K | 2889 | hCoV-19/Australia/VIC13608/2020\|EPI_ISL_563783\|202 | .................... | .................... | ..................... | .................... | .................... | .................... |
| K | 2890 | hCoV-19/Australia/VIC13611/2020\|EPI_ISL_563784\|202 | .................... | .................... | ..................... | .................... | .................... | .................... |
| K | 2891 | hCoV-19/Australia/VIC13613/2020\|EPI_ISL_563785\|202 | .................... | .................... | ..................... | .................... | .................... | .................... |
| K | 2892 | hCoV-19/Australia/VIC13617/2020\|EPI_ISL_563786\|202 | .................... | .................... | ..................... | .................... | .................... | .................... |
| K | 2893 | hCoV-19/Australia/VIC13619/2020\|EPI_ISL_563787\|202 | .................... | .................... | ..................... | .................... | .................... | .................... |
| K | 2894 | hCoV-19/Australia/VIC13622/2020\|EPI_ISL_563788\|202 | .................... | .................... | ..................... | .................... | .................... | .................... |
| K | 2895 | hCoV-19/Australia/VIC13624/2020\|EPI_ISL_563789\|202 | .................... | .................... | ..................... | .................... | .................... | .................... |
| K | 2896 | hCoV-19/Australia/VIC13631/2020\|EPI_ISL_563790\|202 | .................... | .................... | ..................... | .................... | .................... | .................... |
| K | 2897 | hCoV-19/Australia/VIC13635/2020\|EPI_ISL_563791\|202 | .................... | .................... | ..................... | .................... | .................... | .................... |
| K | 2898 | hCoV-19/Australia/VIC13639/2020\|EPI_ISL_563792\|202 | .................... | .................... | ..................... | .................... | .................... | .................... |
| K | 2899 | hCoV-19/Australia/VIC13641/2020\|EPI_ISL_563793\|202 | .................... | .................... | ..................... | .................... | .................... | .................... |
| K | 2900 | hCoV-19/Australia/VIC13642/2020\|EPI_ISL_563794\|202 | .................... | .................... | ..................... | .................... | .................... | .................... |
| K | 2901 | hCoV-19/Australia/VIC13648/2020\|EPI_ISL_563795\|202 | .................... | .................... | ..................... | .................... | .................... | .................... |
| K | 2902 | hCoV-19/Australia/VIC8208/2020\|EPI_ISL_564900\|2020 | .................... | .................... | ..................... | .................... | .................... | .................... |
| K | 2903 | hCoV-19/Australia/VIC8209/2020\|EPI_ISL_564901\|2020 | .................... | .................... | ..................... | .................... | .................... | .................... |
| K | 2904 | hCoV-19/Australia/VIC8210/2020\|EPI_ISL_564902\|2020 | .................... | .................... | ..................... | .................... | .................... | .................... |
| K | 2905 | hCoV-19/Australia/VIC8212/2020\|EPI_ISL_564903\|2020 | .................... | .................... | ..................... | .................... | .................... | .................... |
| K | 2906 | hCoV-19/Australia/VIC8214/2020\|EPI_ISL_564904\|2020 | .................... | .................... | ..................... | .................... | .................... | .................... |
| K | 2907 | hCoV-19/Australia/VIC8215/2020\|EPI_ISL_564905\|2020 | .................... | .................... | ..................... | .................... | .................... | .................... |
| K | 2908 | hCoV-19/Australia/VIC8219/2020\|EPI_ISL_564906\|2020 | .................... | .................... | ..................... | .................... | .................... | .................... |
| K | 2909 | hCoV-19/Australia/VIC8221/2020\|EPI_ISL_564907\|2020 | .................... | .................... | ..................... | .................... | .................... | .................... |
| K | 2910 | hCoV-19/Australia/VIC8222/2020\|EPI_ISL_564908\|2020 | .................... | .................... | ..................... | .................... | .................... | .................... |
| K | 2911 | hCoV-19/Australia/VIC8227/2020\|EPI_ISL_564909\|2020 | .................... | .................... | ..................... | .................... | .................... | .................... |
| K | 2912 | hCoV-19/Australia/VIC8229/2020\|EPI_ISL_564910\|2020 | .................... | .................... | ..................... | .................... | .................... | .................... |
| K | 2913 | hCoV-19/Australia/VIC8232/2020\|EPI_ISL_564911\|2020 | .................... | .................... | ..................... | .................... | .................... | .................... |
| K | 2914 | hCoV-19/Australia/VIC8236/2020\|EPI_ISL_564912\|2020 | .................... | .................... | ..................... | .................... | .................... | .................... |
| K | 2915 | hCoV-19/Australia/VIC8238/2020\|EPI_ISL_564913\|2020 | .................... | .................... | ..................... | .................... | .................... | .................... |
| K | 2916 | hCoV-19/Australia/VIC8239/2020\|EPI_ISL_564914\|2020 | .................... | .................... | ..................... | .................... | .................... | .................... |
| K | 2917 | hCoV-19/Australia/VIC8240/2020\|EPI_ISL_564915\|2020 | .................... | .................... | ..................... | .................... | .................... | .................... |
| K | 2918 | hCoV-19/Australia/VIC8241/2020\|EPI_ISL_564916\|2020 | .................... | .................... | ..................... | .................... | .................... | .................... |
| K | 2919 | hCoV-19/Australia/VIC8243/2020\|EPI_ISL_564917\|2020 | .................... | .................... | ..................... | .................... | .................... | .................... |
| K | 2920 | hCoV-19/Australia/VIC8246/2020\|EPI_ISL_564918\|2020 | .................... | .................... | ..................... | .................... | .................... | .................... |
| K | 2921 | hCoV-19/Australia/VIC8248/2020\|EPI_ISL_564919\|2020 | .................... | .................... | ..................... | .................... | .................... | .................... |
| K | 2922 | hCoV-19/England/QEUH-9C8FE0/2020\|EPI_ISL_567118\|20 | .................... | .................... | ..................... | .................... | .................... | .................... |
| K | 2923 | hCoV-19/England/QEUH-9C83D5/2020\|EPI_ISL_567119\|20 | .................... | .................... | ..................... | .................... | .................... | .................... |

| | | | | | | | | |
|---|---|---|---|---|---|---|---|---|
| K | 2924 | hCoV-19/England/QEUH-9C83E4/2020\|EPI_ISL_567120\|20 | ···················· | ···················· | ····················· | ···················· | ···················· | ···················· |
| K | 2925 | hCoV-19/England/QEUH-9C7F78/2020\|EPI_ISL_567121\|20 | ···················· | ···················· | ····················· | ···················· | ···················· | ···················· |
| K | 2926 | hCoV-19/England/QEUH-9C83B7/2020\|EPI_ISL_567122\|20 | ···················· | ···················· | ····················· | ···················· | ···················· | ···················· |
| K | 2927 | hCoV-19/England/QEUH-9C7BCF/2020\|EPI_ISL_567123\|20 | ···················· | ···················· | ····················· | ···················· | ···················· | ···················· |
| K | 2928 | hCoV-19/England/QEUH-9C8AEF/2020\|EPI_ISL_567124\|20 | ···················· | ···················· | ····················· | ···················· | ···················· | ···················· |
| K | 2929 | hCoV-19/England/QEUH-9C8029/2020\|EPI_ISL_567125\|20 | ···················· | ···················· | ····················· | ···················· | ···················· | ···················· |
| K | 2930 | hCoV-19/England/QEUH-9C85DF/2020\|EPI_ISL_567127\|20 | ···················· | ···················· | ····················· | ···················· | ···················· | ···················· |
| K | 2931 | hCoV-19/England/QEUH-9C84F0/2020\|EPI_ISL_567129\|20 | ···················· | ···················· | ····················· | ···················· | ···················· | ···················· |
| K | 2932 | hCoV-19/England/QEUH-9C90DD/2020\|EPI_ISL_567131\|20 | ···················· | ···················· | ····················· | ···················· | ···················· | ···················· |
| K | 2933 | hCoV-19/England/QEUH-9C8618/2020\|EPI_ISL_567132\|20 | ···················· | ·A·················· | ····················· | ···················· | ···················· | ···A················ |
| K | 2934 | hCoV-19/England/QEUH-9C884F/2020\|EPI_ISL_567133\|20 | ···················· | ···················· | ····················· | ···················· | ···················· | ···················· |
| K | 2935 | hCoV-19/England/QEUH-9C84A5/2020\|EPI_ISL_567135\|20 | ···················· | ···················· | ····················· | ···················· | ···················· | ···················· |
| K | 2936 | hCoV-19/England/QEUH-9C88D6/2020\|EPI_ISL_567137\|20 | ···················· | ···················· | ····················· | ···················· | ···················· | ···················· |
| K | 2937 | hCoV-19/England/QEUH-9C80FC/2020\|EPI_ISL_567140\|20 | ···················· | ···················· | ····················· | ···················· | ···················· | ···················· |
| K | 2938 | hCoV-19/England/QEUH-9C8C43/2020\|EPI_ISL_567141\|20 | ···················· | ···················· | ····················· | ···················· | ···················· | ···················· |
| K | 2939 | hCoV-19/England/QEUH-9C8F4A/2020\|EPI_ISL_567142\|20 | ···················· | ·A·················· | ····················· | ···················· | ···················· | ···················· |
| K | 2940 | hCoV-19/England/QEUH-9C88E5/2020\|EPI_ISL_567143\|20 | ···················· | ·A·················· | ····················· | ···················· | ···················· | ···················· |
| K | 2941 | hCoV-19/England/QEUH-9C8223/2020\|EPI_ISL_567144\|20 | ···················· | ···················· | ····················· | ···················· | ···················· | ···················· |
| K | 2942 | hCoV-19/England/QEUH-9C8C16/2020\|EPI_ISL_567145\|20 | ···················· | ···················· | ····················· | ···················· | ···················· | ···················· |
| K | 2943 | hCoV-19/England/QEUH-9C843C/2020\|EPI_ISL_567146\|20 | ···················· | ···················· | ····················· | ···················· | ···················· | ···················· |
| K | 2944 | hCoV-19/England/QEUH-9C800B/2020\|EPI_ISL_567147\|20 | ···················· | ···················· | ····················· | ···················· | ···················· | ···················· |
| K | 2945 | hCoV-19/England/QEUH-9C8144/2020\|EPI_ISL_567148\|20 | ···················· | ···················· | ····················· | ···················· | ···················· | ···················· |
| K | 2946 | hCoV-19/England/QEUH-9C87D9/2020\|EPI_ISL_567150\|20 | ···················· | ···················· | ····················· | ···················· | ···················· | ···················· |
| K | 2947 | hCoV-19/England/QEUH-9C7AD1/2020\|EPI_ISL_567151\|20 | ···················· | ···················· | ····················· | ···················· | ···················· | ···················· |
| K | 2948 | hCoV-19/England/QEUH-9C8478/2020\|EPI_ISL_567152\|20 | ···················· | ···················· | ····················· | ···················· | ···················· | ···················· |
| K | 2949 | hCoV-19/England/QEUH-9C8CCB/2020\|EPI_ISL_567153\|20 | ···················· | ···················· | ····················· | ···················· | ······T············· | ···················· |
| K | 2950 | hCoV-19/England/QEUH-9C7E8A/2020\|EPI_ISL_567154\|20 | ···················· | ···················· | ····················· | ···················· | ···················· | ···················· |
| K | 2951 | hCoV-19/England/QEUH-9C87BB/2020\|EPI_ISL_567155\|20 | ···················· | ···················· | ····················· | ···················· | ···················· | ···················· |
| K | 2952 | hCoV-19/Wales/QEUH-9C7FD2/2020\|EPI_ISL_567156\|2020 | ···················· | ···················· | ····················· | ···················· | ···················· | ···················· |
| K | 2953 | hCoV-19/England/QEUH-9C9107/2020\|EPI_ISL_567157\|20 | ···················· | ···················· | ····················· | ···················· | ···················· | ···················· |
| K | 2954 | hCoV-19/England/QEUH-9C8DE6/2020\|EPI_ISL_567159\|20 | ···················· | ···················· | ····················· | ···················· | ···················· | ···················· |
| K | 2955 | hCoV-19/England/QEUH-9C7AFF/2020\|EPI_ISL_567160\|20 | ···················· | ···················· | ····················· | ···················· | ···················· | ···················· |
| K | 2956 | hCoV-19/England/QEUH-9C8CF8/2020\|EPI_ISL_567161\|20 | ···················· | ···················· | ····················· | ···················· | ···················· | ···················· |
| K | 2957 | hCoV-19/England/QEUH-9C8D5F/2020\|EPI_ISL_567162\|20 | ···················· | ···················· | ····················· | ···················· | ···················· | ···················· |
| K | 2958 | hCoV-19/England/QEUH-9C7B0B/2020\|EPI_ISL_567164\|20 | ···················· | ···················· | ····················· | ···················· | ···················· | ···················· |
| K | 2959 | hCoV-19/England/QEUH-9C819F/2020\|EPI_ISL_567165\|20 | ···················· | ·A·················· | ····················· | ···················· | ···················· | ···················· |
| K | 2960 | hCoV-19/England/QEUH-9C7F3C/2020\|EPI_ISL_567166\|20 | ···················· | ···················· | ····················· | ···················· | ···················· | ···················· |
| K | 2961 | hCoV-19/England/QEUH-9C87CA/2020\|EPI_ISL_567167\|20 | ···················· | ···················· | ····················· | ···················· | ···················· | ···················· |
| K | 2962 | hCoV-19/England/QEUH-9C7F69/2020\|EPI_ISL_567168\|20 | ···················· | ···················· | ····················· | ···················· | ···················· | ···················· |
| K | 2963 | hCoV-19/England/QEUH-9C8C07/2020\|EPI_ISL_567169\|20 | ···················· | ·A·················· | ····················· | ···················· | ···················· | ···················· |
| K | 2964 | hCoV-19/England/QEUH-9C889A/2020\|EPI_ISL_567171\|20 | ···················· | ···················· | ····················· | ···················· | ···················· | ···················· |
| K | 2965 | hCoV-19/England/QEUH-9C7F87/2020\|EPI_ISL_567172\|20 | ···················· | ···················· | ····················· | ···················· | ···················· | ···················· |
| K | 2966 | hCoV-19/England/QEUH-9C8092/2020\|EPI_ISL_567173\|20 | ···················· | ···················· | ····················· | ···················· | ···················· | ···················· |
| K | 2967 | hCoV-19/England/QEUH-9C82BA/2020\|EPI_ISL_567174\|20 | ···················· | ·A·················· | ····················· | ···················· | ···················· | ···················· |
| K | 2968 | hCoV-19/England/QEUH-9C8A1C/2020\|EPI_ISL_567175\|20 | ···················· | ·A·······A·········· | ····················· | ···················· | ···················· | ···················· |
| K | 2969 | hCoV-19/England/QEUH-9C8CDA/2020\|EPI_ISL_567176\|20 | ···················· | ···················· | ····················· | ···················· | ······T············· | ···················· |
| K | 2970 | hCoV-19/England/QEUH-9C8B37/2020\|EPI_ISL_567177\|20 | ···················· | ···················· | ····················· | ···················· | ···················· | ···················· |
| K | 2971 | hCoV-19/England/QEUH-9C9204/2020\|EPI_ISL_567178\|20 | ···················· | ···················· | ····················· | ···················· | ···················· | ···················· |
| K | 2972 | hCoV-19/England/QEUH-9C7C62/2020\|EPI_ISL_567179\|20 | ···················· | ···················· | ····················· | ···················· | ···················· | ···················· |
| K | 2973 | hCoV-19/England/QEUH-9C7BDE/2020\|EPI_ISL_567180\|20 | ···················· | ····A··············· | ····················· | ···················· | ···················· | ···················· |
| K | 2974 | hCoV-19/England/QEUH-9C7E20/2020\|EPI_ISL_567181\|20 | ···················· | ···················· | ····················· | ···················· | ···················· | ···················· |
| K | 2975 | hCoV-19/England/QEUH-9C834E/2020\|EPI_ISL_567182\|20 | ···················· | ···················· | ····················· | ···················· | ···················· | ···················· |
| K | 2976 | hCoV-19/England/QEUH-9C7EC6/2020\|EPI_ISL_567183\|20 | ···················· | ···················· | ····················· | ···················· | ···················· | ···················· |
| K | 2977 | hCoV-19/Wales/QEUH-9C7EB7/2020\|EPI_ISL_567185\|2020 | ···················· | ···················· | ····················· | ···················· | ···················· | ···················· |
| K | 2978 | hCoV-19/England/QEUH-9C8C8F/2020\|EPI_ISL_567186\|20 | ···················· | ···················· | ····················· | ···················· | ···················· | ···················· |
| K | 2979 | hCoV-19/England/QEUH-9C8BA0/2020\|EPI_ISL_567187\|20 | ···················· | ···················· | ····················· | ···················· | ···················· | ···················· |
| K | 2980 | hCoV-19/England/QEUH-9C7E99/2020\|EPI_ISL_567188\|20 | ···················· | ···················· | ····················· | ···················· | ····T··············· | ···················· |
| K | 2981 | hCoV-19/England/QEUH-9C7C71/2020\|EPI_ISL_567189\|20 | ···················· | ···················· | ····················· | ···················· | ······T············· | ···················· |
| K | 2982 | hCoV-19/England/QEUH-9C9134/2020\|EPI_ISL_567190\|20 | ···················· | ···················· | ····················· | ···················· | ···················· | ···················· |
| K | 2983 | hCoV-19/England/QEUH-9C8232/2020\|EPI_ISL_567191\|20 | ···················· | ···················· | ····················· | ···················· | ···················· | ···················· |
| K | 2984 | hCoV-19/England/QEUH-9C8988/2020\|EPI_ISL_567192\|20 | ···················· | ···················· | ····················· | ···················· | ···················· | ···················· |
| K | 2985 | hCoV-19/England/QEUH-9C8153/2020\|EPI_ISL_567193\|20 | ···················· | ···················· | ····················· | ···················· | ···················· | ···················· |
| K | 2986 | hCoV-19/England/QEUH-9C8117/2020\|EPI_ISL_567194\|20 | ···················· | ···················· | ····················· | ···················· | ···················· | ···················· |
| K | 2987 | hCoV-19/England/QEUH-9C9213/2020\|EPI_ISL_567195\|20 | ···················· | ·A·················· | ····················· | ···················· | ···················· | ···A················ |
| K | 2988 | hCoV-19/England/QEUH-9C8250/2020\|EPI_ISL_567196\|20 | ···················· | ···················· | ····················· | ···················· | ····T··············· | ···················· |
| K | 2989 | hCoV-19/England/QEUH-9C894C/2020\|EPI_ISL_567197\|20 | ···················· | ···················· | ····················· | ···················· | ···················· | ···················· |
| K | 2990 | hCoV-19/England/QEUH-9C7E4E/2020\|EPI_ISL_567198\|20 | ···················· | ···················· | ····················· | ···················· | ···················· | ···················· |
| K | 2991 | hCoV-19/England/QEUH-9C8496/2020\|EPI_ISL_567199\|20 | ···················· | ···················· | ····················· | ···················· | ···················· | ···················· |
| K | 2992 | hCoV-19/Scotland/QEUH-9700F3/2020\|EPI_ISL_568403\|2 | ···················· | ···················· | ····················· | ···················· | ···················· | ···················· |
| K | 2993 | hCoV-19/Scotland/QEUH-96F6B9/2020\|EPI_ISL_568404\|2 | ···················· | ···················· | ····················· | ···················· | ···················· | ···················· |
| K | 2994 | hCoV-19/Scotland/QEUH-98F38F/2020\|EPI_ISL_568449\|2 | ···················· | ···················· | ····················· | ···················· | ···················· | ···················· |
| K | 2995 | hCoV-19/Scotland/QEUH-98EE61/2020\|EPI_ISL_568450\|2 | ···················· | ···················· | ····················· | ···················· | ···················· | ···················· |
| K | 2996 | hCoV-19/Scotland/QEUH-98EE25/2020\|EPI_ISL_568451\|2 | ···················· | ···················· | ····················· | ···················· | ···················· | ···················· |
| K | 2997 | hCoV-19/Scotland/CAMC-99093E/2020\|EPI_ISL_568453\|2 | ···················· | ···················· | ····················· | ···················· | ···················· | ···················· |
| K | 2998 | hCoV-19/Scotland/CAMC-990FF0/2020\|EPI_ISL_568454\|2 | ···················· | ···················· | ····················· | ···················· | ···················· | ···················· |
| K | 2999 | hCoV-19/Scotland/QEUH-98F03D/2020\|EPI_ISL_568455\|2 | ···················· | ···················· | ····················· | ···················· | ···················· | ···················· |
| K | 3000 | hCoV-19/Scotland/QEUH-98F7ED/2020\|EPI_ISL_568456\|2 | ···················· | ···················· | ····················· | ···················· | ···················· | ···················· |
| K | 3001 | hCoV-19/Scotland/QEUH-98F57A/2020\|EPI_ISL_568457\|2 | ···················· | ···················· | ····················· | ···················· | ···················· | ···················· |
| K | 3002 | hCoV-19/Scotland/CAMC-990FE1/2020\|EPI_ISL_568458\|2 | ···················· | ···················· | ····················· | ···················· | ···················· | ···················· |
| K | 3003 | hCoV-19/Scotland/QEUH-98F10D/2020\|EPI_ISL_568459\|2 | ···················· | ···················· | ····················· | ···················· | ···················· | ···················· |

| | | | | | | | | | |
|---|---|---|---|---|---|---|---|---|---|
| K | 3004 | hCoV-19/Scotland/CAMC-990AC2/2020\|EPI_ISL_568461\|2 | .................... | .................... | .................... | .................... | .................... | .................... | |
| K | 3005 | hCoV-19/Scotland/QEUH-98F55C/2020\|EPI_ISL_568462\|2 | .................... | .................... | .................... | .................... | .................... | .................... | |
| K | 3006 | hCoV-19/Scotland/QEUH-98F39E/2020\|EPI_ISL_568463\|2 | .................... | .................... | .................... | .................... | .................... | .................... | |
| K | 3007 | hCoV-19/Scotland/QEUH-98F185/2020\|EPI_ISL_568464\|2 | .................... | .................... | .................... | .................... | .................... | .................... | |
| K | 3008 | hCoV-19/Scotland/ALDP-9128CE/2020\|EPI_ISL_568467\|2 | .................... | ...................A | .................... | .................... | .................... | .................... | 1 |
| K | 3009 | hCoV-19/Scotland/MILK-95999B/2020\|EPI_ISL_568470\|2 | .................... | .................... | .................... | .................... | .................... | .................... | |
| K | 3010 | hCoV-19/Scotland/MILK-95D9E2/2020\|EPI_ISL_568471\|2 | .................... | ...................A | .................... | .................... | .................... | .................... | 1 |
| K | 3011 | hCoV-19/Scotland/CAMC-94907B/2020\|EPI_ISL_568473\|2 | .................... | .................... | .................... | .................... | .................... | .................... | |
| K | 3012 | hCoV-19/Scotland/MILK-5933AA/2020\|EPI_ISL_568476\|2 | .................... | .................... | .................... | .................... | .................... | .................... | |
| K | 3013 | hCoV-19/Russia/OMS-ORINFI-3027S/2020\|EPI_ISL_56978 | .................... | .................... | .................... | .................... | .................... | .................... | |
| K | 3014 | hCoV-19/Russia/OMS-ORINFI-3868S/2020\|EPI_ISL_56978 | .................... | .................... | .................... | .................... | .................... | .................... | |
| K | 3015 | hCoV-19/Russia/OMS-ORINFI-4361S/2020\|EPI_ISL_56978 | .................... | .................... | .................... | .................... | .................... | .................... | |
| K | 3016 | hCoV-19/Russia/OMS-ORINFI-4761S/2020\|EPI_ISL_56978 | .................... | .................... | .................... | .................... | .................... | .................... | |
| K | 3017 | hCoV-19/Russia/OMS-ORINFI-4832S/2020\|EPI_ISL_56978 | .................... | .................... | .................... | .................... | .................... | .................... | |
| K | 3018 | hCoV-19/Russia/OMS-ORINFI-5042S/2020\|EPI_ISL_56978 | .................... | .................... | .................... | .................... | .................... | .................... | |
| K | 3019 | hCoV-19/Russia/OMS-ORINFI-5207S/2020\|EPI_ISL_56978 | .................... | .................... | .................... | .................... | .................... | .................... | |
| K | 3020 | hCoV-19/Russia/OMS-ORINFI-5762S/2020\|EPI_ISL_56978 | .................... | .................... | .................... | .................... | .................... | .................... | |
| K | 3021 | hCoV-19/Russia/OMS-ORINFI-5463S/2020\|EPI_ISL_56978 | .................... | .................... | .................... | .................... | .................... | .................... | |
| K | 3022 | hCoV-19/Russia/OMS-ORINFI-7899S/2020\|EPI_ISL_56978 | .................... | .................... | .................... | ............A....... | .................... | .................... | |
| K | 3023 | hCoV-19/Russia/OMS-ORINFI-8583S/2020\|EPI_ISL_56979 | .................... | .................... | .................... | .................... | .................... | .................... | |
| K | 3024 | hCoV-19/Russia/OMS-ORINFI-8803S/2020\|EPI_ISL_56979 | .................... | .................... | .................... | .................... | .................... | .................... | |
| K | 3025 | hCoV-19/Russia/OMS-ORINFI-8945S/2020\|EPI_ISL_56979 | .................... | .................... | .................... | .................... | .................... | .................... | |
| K | 3026 | hCoV-19/Russia/OMS-ORINFI-9295S/2020\|EPI_ISL_56979 | .................... | .................... | .................... | .................... | .................... | .................... | |
| K | 3027 | hCoV-19/Russia/OMS-ORINFI-9642S/2020\|EPI_ISL_56979 | .................... | .................... | .................... | .................... | .................... | .................... | |
| K | 3028 | hCoV-19/Russia/OMS-ORINFI-9674S/2020\|EPI_ISL_56979 | .................... | .................... | .................... | .................... | .................... | .................... | |
| K | 3029 | hCoV-19/Russia/OMS-ORINFI-4764S/2020\|EPI_ISL_56979 | .................... | .................... | .................... | .................... | .................... | .................... | |
| K | 3030 | hCoV-19/Russia/OMS-ORINFI-4760S/2020\|EPI_ISL_56979 | .................... | .................... | .................... | .................... | .................... | .................... | |
| K | 3031 | hCoV-19/Russia/OMS-ORINFI-4370S/2020\|EPI_ISL_56979 | .................... | .................... | .................... | .................... | .................... | .................... | |
| K | 3032 | hCoV-19/Russia/OMS-ORINFI-1322S/2020\|EPI_ISL_56979 | .................... | .................... | .................... | .................... | .................... | .................... | |
| K | 3033 | hCoV-19/USA/WA-UW-22757/2020\|EPI_ISL_570712\|2020-0 | .................... | .A.................. | .................... | .................... | .................... | .................... | |
| K | 3034 | hCoV-19/USA/WA-UW-22758/2020\|EPI_ISL_570713\|2020-0 | .................... | .................... | .................... | .................... | .................... | .................... | |
| K | 3035 | hCoV-19/USA/WA-UW-23756/2020\|EPI_ISL_570760\|2020-0 | .................... | .A.................. | .................... | .................... | .................... | .................... | |
| K | 3036 | hCoV-19/USA/WA-UW-23804/2020\|EPI_ISL_570761\|2020-0 | .................... | .A.................. | .................... | .................... | .................... | .................... | |
| K | 3037 | hCoV-19/USA/WA-UW-24020/2020\|EPI_ISL_570762\|2020-0 | .................... | .A.................. | .................... | .................... | .................... | .................... | |
| K | 3038 | hCoV-19/USA/WA-UW-24191/2020\|EPI_ISL_570763\|2020-0 | .................... | .................... | .................... | .................... | .................... | .................... | |
| K | 3039 | hCoV-19/USA/WA-UW-24373/2020\|EPI_ISL_570764\|2020-0 | .................... | .................... | .................... | .................... | .................... | .................... | |
| K | 3040 | hCoV-19/USA/WA-UW-246/2020\|EPI_ISL_570765\|2020-03- | .................... | .................... | .................... | .................... | .................... | .................... | |
| K | 3041 | hCoV-19/USA/WA-UW-24736/2020\|EPI_ISL_570766\|2020-0 | .................... | .................... | .................... | .................... | .................... | .................... | |
| K | 3042 | hCoV-19/USA/WA-UW-24775/2020\|EPI_ISL_570767\|2020-0 | .................... | .................... | .................... | .................... | .................... | .................... | |
| K | 3043 | hCoV-19/USA/WA-UW-24787/2020\|EPI_ISL_570768\|2020-0 | .................... | .................... | .................... | .................... | .................... | .................... | |
| K | 3044 | hCoV-19/USA/WA-UW-24839/2020\|EPI_ISL_570769\|2020-0 | .................... | .................... | .................... | .................... | .................... | .................... | |
| K | 3045 | hCoV-19/USA/WA-UW-24901/2020\|EPI_ISL_570770\|2020-0 | .................... | .................... | .................... | .................... | .................... | .................... | |
| K | 3046 | hCoV-19/USA/WA-UW-24906/2020\|EPI_ISL_570771\|2020-0 | .................... | .................... | .................... | .................... | .................... | .................... | |
| K | 3047 | hCoV-19/USA/WA-UW-24925/2020\|EPI_ISL_570772\|2020-0 | .................... | .A.................. | .................... | .................... | .................... | .................... | |
| K | 3048 | hCoV-19/USA/WA-UW-24930/2020\|EPI_ISL_570773\|2020-0 | .................... | .A.................. | .................... | .................... | .................... | .................... | |
| K | 3049 | hCoV-19/USA/WA-UW-24935/2020\|EPI_ISL_570774\|2020-0 | .................... | .................... | .................... | .................... | .................... | .................... | |
| K | 3050 | hCoV-19/USA/WA-UW-24937/2020\|EPI_ISL_570775\|2020-0 | .................... | .................... | .................... | .................... | .................... | .................... | |
| K | 3051 | hCoV-19/USA/WA-UW-24941/2020\|EPI_ISL_570776\|2020-0 | .................... | .................... | .................... | .................... | .................... | .................... | |
| K | 3052 | hCoV-19/USA/WA-UW-24954/2020\|EPI_ISL_570777\|2020-0 | .................... | .................... | .................... | .................... | .................... | .................... | |
| K | 3053 | hCoV-19/USA/WA-UW-26488/2020\|EPI_ISL_570778\|2020-0 | .................... | .................... | .................... | .................... | .................... | .................... | |
| K | 3054 | hCoV-19/USA/WA-UW-26849/2020\|EPI_ISL_570779\|2020-0 | .................... | .A.................. | .................... | .................... | .................... | .................... | |
| K | 3055 | hCoV-19/USA/WA-UW-26858/2020\|EPI_ISL_570780\|2020-0 | .................... | .................... | .................... | .................... | .................... | .................... | |
| K | 3056 | hCoV-19/USA/WA-UW-26859/2020\|EPI_ISL_570781\|2020-0 | .................... | .A.................. | .................... | .................... | .................... | .................... | |
| K | 3057 | hCoV-19/USA/WA-UW-26860/2020\|EPI_ISL_570782\|2020-0 | .................... | .................... | .................... | .................... | .................... | .................... | |
| K | 3058 | hCoV-19/USA/WA-UW-26861/2020\|EPI_ISL_570783\|2020-0 | .................... | .................... | .................... | .................... | .................... | .................... | |
| K | 3059 | hCoV-19/USA/WA-UW-26862/2020\|EPI_ISL_570784\|2020-0 | .................... | .................... | .................... | .................... | .................... | .................... | |
| K | 3060 | hCoV-19/USA/WA-UW-26867/2020\|EPI_ISL_570785\|2020-0 | .................... | .................... | .................... | .................... | .................... | .................... | |
| K | 3061 | hCoV-19/USA/WA-UW-26868/2020\|EPI_ISL_570786\|2020-0 | .................... | .............A...... | .................... | .................... | .................... | .................... | |
| K | 3062 | hCoV-19/USA/WA-UW-27032/2020\|EPI_ISL_570787\|2020-0 | .................... | .A.................. | .................... | .................... | .................... | .................... | |
| K | 3063 | hCoV-19/USA/WA-UW-27033/2020\|EPI_ISL_570788\|2020-0 | .................... | .................... | .................... | .................... | .................... | .................... | |
| K | 3064 | hCoV-19/USA/WA-UW-27034/2020\|EPI_ISL_570789\|2020-0 | .................... | .................... | .................... | .................... | .................... | .................... | |
| K | 3065 | hCoV-19/USA/WA-UW-27038/2020\|EPI_ISL_570790\|2020-0 | .................... | .................... | .................... | .................... | .................... | .................... | |
| K | 3066 | hCoV-19/USA/WA-UW-27042/2020\|EPI_ISL_570791\|2020-0 | .................... | .A.................. | .................... | .................... | .................... | .................... | |
| K | 3067 | hCoV-19/USA/WA-UW-27044/2020\|EPI_ISL_570792\|2020-0 | .................... | .A.................. | .................... | .................... | .................... | .................... | |
| K | 3068 | hCoV-19/USA/WA-UW-27046/2020\|EPI_ISL_570793\|2020-0 | .................... | .................... | .................... | .................... | .................... | .................... | |
| K | 3069 | hCoV-19/USA/WA-UW-27498/2020\|EPI_ISL_570794\|2020-0 | .................... | .A.................. | .................... | .................... | .................... | .................... | |
| K | 3070 | hCoV-19/USA/WA-UW-27499/2020\|EPI_ISL_570795\|2020-0 | .................... | .A.................. | .................... | .................... | .................... | .................... | |
| K | 3071 | hCoV-19/USA/WA-UW-27500/2020\|EPI_ISL_570796\|2020-0 | .................... | .................... | .................... | .................... | .................... | .................... | |
| K | 3072 | hCoV-19/USA/WA-UW-27501/2020\|EPI_ISL_570797\|2020-0 | .................... | .................... | .................... | .................... | .................... | .................... | |
| K | 3073 | hCoV-19/Gambia/3761/2020\|EPI_ISL_561013\|2020-07-30 | .................... | .................... | .................... | .................... | .................... | .................... | |
| K | 3074 | hCoV-19/Gambia/GC19-1503/2020\|EPI_ISL_561021\|2020- | .................... | .................... | .................... | .................... | .................... | .................... | |
| K | 3075 | hCoV-19/Gambia/GC19-1527/2020\|EPI_ISL_561022\|2020- | .................... | .................... | .................... | .................... | .................... | .................... | |
| K | 3076 | hCoV-19/Gambia/GC19-1818/2020\|EPI_ISL_561034\|2020- | .................... | .................... | .................... | .................... | .................... | .................... | |
| K | 3077 | hCoV-19/Gambia/GC19-1983/2020\|EPI_ISL_561038\|2020- | .................... | .................... | .................... | .................... | .................... | .................... | |
| K | 3078 | hCoV-19/Gambia/GC19-2197/2020\|EPI_ISL_561040\|2020- | .................... | .................... | .................... | .................... | .................... | .................... | |
| K | 3079 | hCoV-19/Gambia/GC19-3087/2020\|EPI_ISL_561041\|2020- | .................... | .................... | .................... | .................... | .................... | .................... | |
| K | 3080 | hCoV-19/Gambia/GC19-3140/2020\|EPI_ISL_561042\|2020- | .................... | .................... | .................... | .................... | .................... | .................... | |
| K | 3081 | hCoV-19/Gambia/GC19-3165/2020\|EPI_ISL_561043\|2020- | .................... | .................... | .................... | .................... | .................... | .................... | |
| K | 3088 | hCoV-19/Gambia/GC19-3219/2020\|EPI_ISL_561052\|2020- | .................... | .................... | .................... | .................... | .................... | .................... | |
| K | 3095 | hCoV-19/Gambia/GC19-3317/2020\|EPI_ISL_561060\|2020- | .................... | .................... | .................... | .................... | .................... | .................... | |

| | | | | | | | | |
|---|---|---|---|---|---|---|---|---|
| K | 3097 | hCoV-19/Gambia/GC19-3358/2020\|EPI_ISL_561062\|2020- | ···················· | ···················· | ···················· | ···················· | ···················· | ···················· |
| K | 3103 | hCoV-19/Gambia/GC19-3381/2020\|EPI_ISL_561068\|2020- | ···················· | ···················· | ···················· | ···················· | ···················· | ···················· |
| K | 3104 | hCoV-19/Gambia/GC19-3396/2020\|EPI_ISL_561069\|2020- | ···················· | ···················· | ···················· | ···················· | ···················· | ···················· |
| K | 3105 | hCoV-19/Gambia/GC19-3398/2020\|EPI_ISL_561070\|2020- | ···················· | ···················· | ···················· | ···················· | ···················· | ···················· |
| K | 3106 | hCoV-19/Gambia/GC19-3768/2020\|EPI_ISL_561071\|2020- | ···················· | ···················· | ···················· | ···················· | ···················· | ···················· |
| K | 3107 | hCoV-19/Gambia/GC19-3773/2020\|EPI_ISL_561073\|2020- | ···················· | ···················· | ···················· | ···················· | ···················· | ···················· |
| K | 3108 | hCoV-19/Gambia/GC19-3774/2020\|EPI_ISL_561074\|2020- | ···················· | ···················· | ···················· | ···················· | ···················· | ···················· |
| K | 3109 | hCoV-19/Gambia/GC19-3775/2020\|EPI_ISL_561075\|2020- | ···················· | ···················· | ···················· | ···················· | ···················· | ···················· |
| K | 3110 | hCoV-19/Gambia/GC19-3776/2020\|EPI_ISL_561076\|2020- | ···················· | ···················· | ···················· | ···················· | ···················· | ···················· |
| K | 3111 | hCoV-19/Gambia/GC19-3792/2020\|EPI_ISL_561077\|2020- | ···················· | ···················· | ···················· | ···················· | ···················· | ···················· |
| K | 3112 | hCoV-19/Gambia/GC19-3796/2020\|EPI_ISL_561079\|2020- | ···················· | ···················· | ···················· | ···················· | ···················· | ···················· |
| K | 3113 | hCoV-19/Gambia/GC19-3798/2020\|EPI_ISL_561080\|2020- | ···················· | ···················· | ···················· | ···················· | ···················· | ···················· |
| K | 3114 | hCoV-19/Gambia/GC19-3799/2020\|EPI_ISL_561081\|2020- | ···················· | ···················· | ···················· | ···················· | ···················· | ···················· |
| K | 3115 | hCoV-19/Gambia/GC19-3802/2020\|EPI_ISL_561082\|2020- | ···················· | ···················· | ···················· | ···················· | ···················· | ···················· |
| K | 3116 | hCoV-19/Gambia/GC19-3805/2020\|EPI_ISL_561083\|2020- | ···················· | ···················· | ···················· | ···················· | ···················· | ···················· |
| K | 3117 | hCoV-19/Gambia/GC19-3806/2020\|EPI_ISL_561084\|2020- | ···················· | ···················· | ···················· | ···················· | ···················· | ···················· |
| K | 3118 | hCoV-19/Gambia/GC19-3807/2020\|EPI_ISL_561085\|2020- | ···················· | ···················· | ···················· | ···················· | ···················· | ···················· |
| K | 3120 | hCoV-19/Gambia/GC19-3813/2020\|EPI_ISL_561087\|2020- | ···················· | ···················· | ···················· | ···················· | ···················· | ···················· |
| K | 3121 | hCoV-19/Gambia/GC19-3814/2020\|EPI_ISL_561088\|2020- | ···················· | ···················· | ···················· | ···················· | ···················· | ···················· |
| K | 3123 | hCoV-19/Gambia/GC19-3818/2020\|EPI_ISL_561090\|2020- | ···················· | ···················· | ···················· | ···················· | ···················· | ···················· |
| K | 3124 | hCoV-19/Gambia/GC19-3820/2020\|EPI_ISL_561091\|2020- | ···················· | ···················· | ···················· | ···················· | ···················· | ···················· |
| K | 3125 | hCoV-19/Gambia/GC19-3822/2020\|EPI_ISL_561093\|2020- | ···················· | ···················· | ···················· | ···················· | ···················· | ···················· |
| K | 3127 | hCoV-19/Gambia/GC19-3831/2020\|EPI_ISL_561095\|2020- | ···················· | ···················· | ···················· | ···················· | ···················· | ···················· |
| K | 3128 | hCoV-19/Gambia/GC19-3835/2020\|EPI_ISL_561096\|2020- | ···················· | ···················· | ···················· | ···················· | ···················· | ···················· |
| K | 3129 | hCoV-19/Gambia/GC19-3836/2020\|EPI_ISL_561097\|2020- | ···················· | ···················· | ···················· | ···················· | ···················· | ···················· |
| K | 3130 | hCoV-19/Gambia/GC19-3839/2020\|EPI_ISL_561098\|2020- | ···················· | ···················· | ···················· | ···················· | ···················· | ···················· |
| K | 3131 | hCoV-19/Gambia/GC19-3842/2020\|EPI_ISL_561099\|2020- | ···················· | ···················· | ···················· | ···················· | ···················· | ···················· |
| K | 3132 | hCoV-19/Australia/VIC12921/2020\|EPI_ISL_563606\|202 | ···················· | ···················· | ···················· | ···················· | ···················· | ···················· |
| K | 3133 | hCoV-19/Australia/VIC13327/2020\|EPI_ISL_563675\|202 | ···················· | ···················· | ···················· | ···················· | ···················· | ···················· |
| K | 3134 | hCoV-19/Australia/VIC13329/2020\|EPI_ISL_563676\|202 | ···················· | ···················· | ···················· | ···················· | ···················· | ···················· |
| K | 3135 | hCoV-19/Australia/VIC13330/2020\|EPI_ISL_563677\|202 | ···················· | ···················· | ···················· | ···················· | ···················· | ···················· |
| K | 3136 | hCoV-19/Australia/VIC13331/2020\|EPI_ISL_563678\|202 | ···················· | ···················· | ···················· | ···················· | ···················· | ···················· |
| K | 3137 | hCoV-19/Australia/VIC13332/2020\|EPI_ISL_563679\|202 | ···················· | ···················· | ···················· | ···················· | ···················· | ···················· |
| K | 3138 | hCoV-19/Australia/VIC13334/2020\|EPI_ISL_563680\|202 | ···················· | ···················· | ···················· | ···················· | ···················· | ···················· |
| K | 3139 | hCoV-19/Australia/VIC13336/2020\|EPI_ISL_563681\|202 | ···················· | ···················· | ···················· | ···················· | ···················· | ···················· |
| K | 3140 | hCoV-19/Australia/VIC13337/2020\|EPI_ISL_563682\|202 | ···················· | ···················· | ···················· | ···················· | N··················· | ···················· |
| K | 3141 | hCoV-19/Australia/VIC13340/2020\|EPI_ISL_563683\|202 | ···················· | ···················· | ···················· | ···················· | ···················· | ···················· |
| K | 3142 | hCoV-19/Australia/VIC13341/2020\|EPI_ISL_563684\|202 | ···················· | ···················· | ···················· | ···················· | ···················· | ···················· |
| K | 3143 | hCoV-19/Australia/VIC13344/2020\|EPI_ISL_563685\|202 | ···················· | ···················· | ···················· | ···················· | ···················· | ···················· |
| K | 3144 | hCoV-19/Australia/VIC13347/2020\|EPI_ISL_563686\|202 | ···················· | ···················· | ···················· | ···················· | ···················· | ···················· |
| K | 3145 | hCoV-19/Australia/VIC13350/2020\|EPI_ISL_563687\|202 | ···················· | ···················· | ···················· | ···················· | ···················· | ···················· |
| K | 3146 | hCoV-19/Australia/VIC13352/2020\|EPI_ISL_563688\|202 | ···················· | ···················· | ···················· | ···················· | ···················· | ···················· |
| K | 3147 | hCoV-19/Australia/VIC13353/2020\|EPI_ISL_563689\|202 | ···················· | ···················· | ···················· | ···················· | ···················· | ···················· |
| K | 3148 | hCoV-19/Australia/VIC13354/2020\|EPI_ISL_563690\|202 | ···················· | ···················· | ···················· | ···················· | ···················· | ···················· |
| K | 3149 | hCoV-19/Australia/VIC13355/2020\|EPI_ISL_563691\|202 | ···················· | ···················· | ···················· | ···················· | ···················· | ···················· |
| K | 3150 | hCoV-19/Australia/VIC13356/2020\|EPI_ISL_563692\|202 | ···················· | ···················· | ···················· | ···················· | ···················· | ···················· |
| K | 3151 | hCoV-19/Australia/VIC13365/2020\|EPI_ISL_563693\|202 | ···················· | ···················· | ···················· | ···················· | ···················· | ···················· |
| K | 3152 | hCoV-19/Australia/VIC13367/2020\|EPI_ISL_563694\|202 | ···················· | ···················· | ···················· | ···················· | ···················· | ···················· |
| K | 3153 | hCoV-19/Australia/VIC13369/2020\|EPI_ISL_563695\|202 | ···················· | ···················· | ···················· | ···················· | ···················· | ···················· |
| K | 3155 | hCoV-19/Australia/VIC13371/2020\|EPI_ISL_563697\|202 | ···················· | ···················· | ···················· | ···················· | ···················· | ···················· |
| K | 3156 | hCoV-19/Australia/VIC13373/2020\|EPI_ISL_563698\|202 | ···················· | ···················· | ···················· | ···················· | ···················· | ···················· |
| K | 3157 | hCoV-19/Australia/VIC13374/2020\|EPI_ISL_563699\|202 | ···················· | ···················· | ···················· | ···················· | ···················· | ···················· |
| K | 3158 | hCoV-19/Australia/VIC8315/2020\|EPI_ISL_564937\|2020 | ···················· | ···················· | ···················· | ···················· | ···················· | ···················· |
| K | 3159 | hCoV-19/Australia/VIC8322/2020\|EPI_ISL_564938\|2020 | ···················· | ···················· | ···················· | ···················· | ···················· | ···················· |
| K | 3160 | hCoV-19/England/QEUH-9C88A9/2020\|EPI_ISL_567100\|20 | ···················· | ···················· | ···················· | ···················· | ······T············· | ···················· |
| K | 3161 | hCoV-19/England/QEUH-9C8636/2020\|EPI_ISL_567103\|20 | ···················· | ···················· | ···················· | ···················· | ···················· | ···················· |
| K | 3162 | hCoV-19/England/QEUH-9C7FF0/2020\|EPI_ISL_567104\|20 | ···················· | ···················· | ···················· | ···················· | ···················· | ···················· |
| K | 3163 | hCoV-19/England/QEUH-9C7E02/2020\|EPI_ISL_567105\|20 | ···················· | ·A·················· | ···················· | ···················· | ···················· | ···················· |
| K | 3164 | hCoV-19/England/QEUH-9C8B64/2020\|EPI_ISL_567106\|20 | ···················· | ···················· | ···················· | ···················· | ···················· | ···················· |
| K | 3165 | hCoV-19/England/QEUH-9C85EE/2020\|EPI_ISL_567107\|20 | ···················· | ···················· | ···················· | ···················· | ···················· | ···················· |
| K | 3166 | hCoV-19/Wales/QEUH-9C8047/2020\|EPI_ISL_567108\|2020 | ···················· | ···················· | ···················· | ···················· | ···················· | ···················· |
| K | 3167 | hCoV-19/England/QEUH-9C7DAB/2020\|EPI_ISL_567109\|20 | ···················· | ···················· | ···················· | ···················· | ···················· | ···················· |
| K | 3168 | hCoV-19/England/QEUH-9C7B74/2020\|EPI_ISL_567111\|20 | ···················· | ···················· | ···················· | ···················· | ···················· | ···················· |
| K | 3169 | hCoV-19/England/QEUH-9C91CB/2020\|EPI_ISL_567112\|20 | ···················· | ···················· | ···················· | ···················· | ···················· | ···················· |
| K | 3170 | hCoV-19/England/QEUH-9C8BEC/2020\|EPI_ISL_567115\|20 | ···················· | ·A·················· | ···················· | ···················· | ···················· | ···A················ |
| K | 3171 | hCoV-19/England/QEUH-9C86BE/2020\|EPI_ISL_567116\|20 | ···················· | ···················· | ···················· | ···················· | ···················· | ···················· |
| K | 3172 | hCoV-19/England/QEUH-9C8979/2020\|EPI_ISL_567117\|20 | ···················· | ···················· | ···················· | ···················· | ···················· | ···················· |
| K | 3173 | hCoV-19/England/QEUH-97012D/2020\|EPI_ISL_568401\|20 | ···················· | ···················· | ···················· | ···················· | ···················· | ···················· |
| K | 3174 | hCoV-19/Scotland/QEUH-96FDF0/2020\|EPI_ISL_568402\|2 | ···················· | ···················· | ···················· | ···················· | ···················· | ···················· |
| K | 3175 | hCoV-19/England/QEUH-97013C/2020\|EPI_ISL_568405\|20 | ···················· | ···················· | ···················· | ···················· | ···················· | ···················· |
| K | 3176 | hCoV-19/Scotland/QEUH-96FB7E/2020\|EPI_ISL_568406\|2 | ···················· | ···················· | ···················· | ···················· | ···················· | ···················· |
| K | 3177 | hCoV-19/Scotland/QEUH-96F5BC/2020\|EPI_ISL_568407\|2 | ···················· | ···················· | ···················· | ···················· | ···················· | ···················· |
| K | 3178 | hCoV-19/Scotland/QEUH-96F69B/2020\|EPI_ISL_568408\|2 | ···················· | ···················· | ···················· | ···················· | ···················· | ···················· |
| K | 3179 | hCoV-19/Scotland/QEUH-96FEEE/2020\|EPI_ISL_568411\|2 | ···················· | ···················· | ···················· | ···················· | ···················· | ···················· |
| K | 3180 | hCoV-19/England/QEUH-97010F/2020\|EPI_ISL_568412\|20 | ···················· | ···················· | ···················· | ···················· | ···················· | ···················· |
| K | 3181 | hCoV-19/Scotland/QEUH-96FE66/2020\|EPI_ISL_568413\|2 | ···················· | ···················· | ···················· | ···················· | ···················· | ···················· |
| K | 3182 | hCoV-19/Scotland/QEUH-96FEC0/2020\|EPI_ISL_568414\|2 | ···················· | ···················· | ···················· | ···················· | ···················· | ···················· |
| K | 3183 | hCoV-19/Scotland/QEUH-96FD78/2020\|EPI_ISL_568416\|2 | ···················· | ···················· | ···················· | ···················· | ···················· | ···················· |
| K | 3184 | hCoV-19/Scotland/QEUH-96FBC9/2020\|EPI_ISL_568417\|2 | ···················· | ···················· | ···················· | ···················· | ···················· | ···················· |
| K | 3185 | hCoV-19/Scotland/QEUH-96F6AA/2020\|EPI_ISL_568418\|2 | ···················· | ···················· | ···················· | ···················· | ···················· | ···················· |

| | | | | | | | | |
|---|---|---|---|---|---|---|---|---|
| K | 3186 | hCoV-19/Scotland/QEUH-96FD69/2020\|EPI_ISL_568420\|2 | ···················· | ···················· | ···················· | ···················· | ···················· | ···················· |
| K | 3187 | hCoV-19/Scotland/QEUH-96FC02/2020\|EPI_ISL_568421\|2 | ···················· | ···················· | ···················· | ···················· | ···················· | ···················· |
| K | 3188 | hCoV-19/Scotland/QEUH-96FD4B/2020\|EPI_ISL_568423\|2 | ···················· | ·A·················· | ···················· | ···················· | ···················· | ···················· |
| K | 3189 | hCoV-19/Scotland/QEUH-96F9FC/2020\|EPI_ISL_568424\|2 | ···················· | ···················· | ···················· | ···················· | ···················· | ···················· |
| K | 3190 | hCoV-19/Scotland/QEUH-96FEB1/2020\|EPI_ISL_568425\|2 | ···················· | ···················· | ···················· | ···················· | ···················· | ···················· |
| K | 3191 | hCoV-19/Scotland/QEUH-96FB05/2020\|EPI_ISL_568426\|2 | ···················· | ···················· | ···················· | ···················· | ···················· | ···················· |
| K | 3192 | hCoV-19/Scotland/QEUH-96FE48/2020\|EPI_ISL_568427\|2 | ···················· | ···················· | ···················· | ···················· | ···················· | ···················· |
| K | 3193 | hCoV-19/Scotland/QEUH-96FC99/2020\|EPI_ISL_568429\|2 | ···················· | ···················· | ···················· | ···················· | ···················· | ···················· |
| K | 3194 | hCoV-19/Scotland/QEUH-96FF45/2020\|EPI_ISL_568430\|2 | ···················· | ···················· | ···················· | ···················· | ···················· | ···················· |
| K | 3195 | hCoV-19/Scotland/QEUH-98F3DA/2020\|EPI_ISL_568431\|2 | ···················· | ···················· | ···················· | ···················· | ···················· | ···················· |
| K | 3196 | hCoV-19/Scotland/QEUH-98F02E/2020\|EPI_ISL_568432\|2 | ···················· | ···················· | ···················· | ···················· | ···················· | ···················· |
| K | 3197 | hCoV-19/Scotland/QEUH-98EFC8/2020\|EPI_ISL_568433\|2 | ···················· | ···················· | ···················· | ···················· | ···················· | ···················· |
| K | 3198 | hCoV-19/Scotland/QEUH-98F079/2020\|EPI_ISL_568434\|2 | ···················· | ···················· | ···················· | ···················· | ···················· | ···················· |
| K | 3199 | hCoV-19/Scotland/CAMC-9907BC/2020\|EPI_ISL_568436\|2 | ···················· | ···················· | ···················· | ···················· | ···················· | ···················· |
| K | 3200 | hCoV-19/Scotland/QEUH-98F53E/2020\|EPI_ISL_568437\|2 | ···················· | ···················· | ···················· | ···················· | ···················· | ···················· |
| K | 3201 | hCoV-19/Scotland/QEUH-98F361/2020\|EPI_ISL_568438\|2 | ···················· | ···················· | ···················· | ···················· | ···················· | ···················· |
| K | 3202 | hCoV-19/Scotland/QEUH-98F5F2/2020\|EPI_ISL_568439\|2 | ···················· | ···················· | ···················· | ···················· | ···················· | ···················· |
| K | 3203 | hCoV-19/Scotland/QEUH-98EE70/2020\|EPI_ISL_568440\|2 | ···················· | ···················· | ···················· | ···················· | ···················· | ···················· |
| K | 3204 | hCoV-19/Scotland/QEUH-98F334/2020\|EPI_ISL_568441\|2 | ···················· | ···················· | ···················· | ···················· | ···················· | ···················· |
| K | 3205 | hCoV-19/Scotland/QEUH-98F219/2020\|EPI_ISL_568442\|2 | ···················· | ···················· | ···················· | ···················· | ···················· | ···················· |
| K | 3206 | hCoV-19/Scotland/QEUH-98F3F8/2020\|EPI_ISL_568443\|2 | ···················· | ···················· | ···················· | ···················· | ···················· | ···················· |
| K | 3207 | hCoV-19/Scotland/QEUH-98EFD7/2020\|EPI_ISL_568444\|2 | ···················· | ···················· | ···················· | ···················· | ···················· | ···················· |
| K | 3208 | hCoV-19/Scotland/QEUH-98F2FB/2020\|EPI_ISL_568445\|2 | ···················· | ···················· | ···················· | ···················· | ···················· | ···················· |
| K | 3209 | hCoV-19/Scotland/QEUH-98F431/2020\|EPI_ISL_568446\|2 | ···················· | ···················· | ···················· | ···················· | ···················· | ···················· |
| K | 3210 | hCoV-19/Scotland/QEUH-98EF31/2020\|EPI_ISL_568447\|2 | ···················· | ···················· | ···················· | ···················· | ···················· | ···················· |
| K | 3211 | hCoV-19/Scotland/QEUH-98F352/2020\|EPI_ISL_568448\|2 | ···················· | ···················· | ···················· | ···················· | ···················· | ···················· |
| K | 3212 | hCoV-19/USA/TX-DSHS-0610/2020\|EPI_ISL_569720\|2020- | ···················· | ···················· | ···················· | ···················· | ···················· | ···················· |
| K | 3213 | hCoV-19/Russia/OMS-ORINFI-5710S/2020\|EPI_ISL_56973 | ···················· | ···················· | ···················· | ···················· | ···················· | ···················· |
| K | 3214 | hCoV-19/Russia/OMS-ORINFI-5819S/2020\|EPI_ISL_56973 | ···················· | ···················· | ···················· | ···················· | ···················· | ···················· |
| K | 3215 | hCoV-19/Russia/OMS-ORINFI-5835S/2020\|EPI_ISL_56973 | ···················· | ···················· | ···················· | ···················· | ···················· | ···················· |
| K | 3216 | hCoV-19/Russia/OMS-ORINFI-6215S/2020\|EPI_ISL_56973 | ···················· | ···················· | ···················· | ···················· | ···················· | ···················· |
| K | 3217 | hCoV-19/Russia/OMS-ORINFI-6673S/2020\|EPI_ISL_56973 | ···················· | ···················· | ···················· | ···················· | ···················· | ···················· |
| K | 3218 | hCoV-19/Russia/OMS-ORINFI-6675S/2020\|EPI_ISL_56974 | ···················· | ···················· | ···················· | ···················· | ···················· | ···················· |
| K | 3219 | hCoV-19/Russia/OMS-ORINFI-6889S/2020\|EPI_ISL_56974 | ···················· | ···················· | ···················· | ···················· | ···················· | ···················· |
| K | 3220 | hCoV-19/Russia/OMS-ORINFI-7121S/2020\|EPI_ISL_56974 | ···················· | ···················· | ···················· | ···················· | ···················· | ···················· |
| K | 3221 | hCoV-19/Russia/OMS-ORINFI-7126S/2020\|EPI_ISL_56974 | ···················· | ···················· | ···················· | ···················· | ···················· | ···················· |
| K | 3222 | hCoV-19/Russia/OMS-ORINFI-7132S/2020\|EPI_ISL_56974 | ···················· | ···················· | ···················· | ···················· | ···················· | ···················· |
| K | 3223 | hCoV-19/Russia/OMS-ORINFI-7146S/2020\|EPI_ISL_56974 | ···················· | ···················· | ···················· | ···················· | ···················· | ···················· |
| K | 3224 | hCoV-19/Russia/OMS-ORINFI-9080S/2020\|EPI_ISL_56974 | ···················· | ···················· | ···················· | ···················· | ···················· | ···················· |
| K | 3225 | hCoV-19/Russia/OMS-ORINFI-9431S/2020\|EPI_ISL_56974 | ···················· | ···················· | ···················· | ···················· | ···················· | ···················· |
| K | 3226 | hCoV-19/Russia/OMS-ORINFI-9433S/2020\|EPI_ISL_56974 | ···················· | ···················· | ···················· | ···················· | ···················· | ···················· |
| K | 3227 | hCoV-19/Russia/OMS-ORINFI-9440S/2020\|EPI_ISL_56974 | ···················· | ···················· | ···················· | ···················· | ···················· | ···················· |
| K | 3228 | hCoV-19/Russia/OMS-ORINFI-9064S/2020\|EPI_ISL_56975 | ···················· | ···················· | ···················· | ···················· | ···················· | ···················· |
| K | 3229 | hCoV-19/Russia/OMS-ORINFI-543S/2020\|EPI_ISL_569751 | ···················· | ···················· | ···················· | ···················· | ···················· | ···················· |
| K | 3230 | hCoV-19/Russia/OMS-ORINFI-1129S/2020\|EPI_ISL_56975 | ···················· | ···················· | ···················· | ···················· | ···················· | ···················· |
| K | 3231 | hCoV-19/Russia/OMS-ORINFI-9761S/2020\|EPI_ISL_56975 | ···················· | ···················· | ···················· | ···················· | ···················· | ···················· |
| K | 3232 | hCoV-19/Russia/OMS-ORINFI-9785S/2020\|EPI_ISL_56975 | ···················· | ···················· | ···················· | ···················· | ···················· | ···················· |
| K | 3233 | hCoV-19/Russia/OMS-ORINFI-9809S/2020\|EPI_ISL_56975 | ···················· | ···················· | ···················· | ···················· | ···················· | ···················· |
| K | 3234 | hCoV-19/Russia/OMS-ORINFI-9815S/2020\|EPI_ISL_56975 | ···················· | ···················· | ···················· | ···················· | ···················· | ···················· |
| K | 3235 | hCoV-19/Russia/OMS-ORINFI-9820S/2020\|EPI_ISL_56975 | ···················· | ···················· | ···················· | ···················· | ···················· | ···················· |
| K | 3236 | hCoV-19/Russia/OMS-ORINFI-1296S/2020\|EPI_ISL_56975 | ···················· | ···················· | ···················· | ···················· | ···················· | ···················· |
| K | 3237 | hCoV-19/Russia/OMS-ORINFI-2022S/2020\|EPI_ISL_56975 | ···················· | ···················· | ···················· | ···················· | ···················· | ···················· |
| K | 3238 | hCoV-19/Russia/OMS-ORINFI-2306S/2020\|EPI_ISL_56976 | ···················· | ···················· | ···················· | ···················· | ···················· | ···················· |
| K | 3239 | hCoV-19/Russia/OMS-ORINFI-2854S/2020\|EPI_ISL_56976 | ···················· | ···················· | ···················· | ···················· | ···················· | ···················· |
| K | 3240 | hCoV-19/Russia/OMS-ORINFI-2990S/2020\|EPI_ISL_56976 | ···················· | ···················· | ···················· | ···················· | ···················· | ···················· |
| K | 3241 | hCoV-19/Russia/OMS-ORINFI-3807S/2020\|EPI_ISL_56976 | ···················· | ···················· | ···················· | ···················· | ···················· | ···················· |
| K | 3242 | hCoV-19/Russia/OMS-ORINFI-3811S/2020\|EPI_ISL_56976 | ···················· | ···················· | ···················· | ···················· | ···················· | ···················· |
| K | 3243 | hCoV-19/Russia/OMS-ORINFI-3846S/2020\|EPI_ISL_56976 | ···················· | ···················· | ···················· | ···················· | ···················· | ···················· |
| K | 3244 | hCoV-19/Russia/OMS-ORINFI-4143S/2020\|EPI_ISL_56976 | ···················· | ···················· | ···················· | ···················· | ···················· | ···················· |
| K | 3245 | hCoV-19/Russia/OMS-ORINFI-4507S/2020\|EPI_ISL_56976 | ···················· | ···················· | ···················· | ···················· | ···················· | ···················· |
| K | 3246 | hCoV-19/Russia/OMS-ORINFI-5350S/2020\|EPI_ISL_56976 | ···················· | ···················· | ···················· | ···················· | ···················· | ···················· |
| K | 3247 | hCoV-19/Russia/OMS-ORINFI-5363S/2020\|EPI_ISL_56976 | ···················· | ···················· | ···················· | ···················· | ···················· | ···················· |
| K | 3248 | hCoV-19/Russia/OMS-ORINFI-5372S/2020\|EPI_ISL_56977 | ···················· | ···················· | ···················· | ···················· | ···················· | ···················· |
| K | 3249 | hCoV-19/Russia/OMS-ORINFI-5782S/2020\|EPI_ISL_56977 | ···················· | ···················· | ···················· | ···················· | ···················· | ···················· |
| K | 3250 | hCoV-19/Russia/OMS-ORINFI-5900S/2020\|EPI_ISL_56977 | ···················· | ···················· | ···················· | ···················· | ···················· | ···················· |
| K | 3251 | hCoV-19/Russia/OMS-ORINFI-5948S/2020\|EPI_ISL_56977 | ···················· | ···················· | ···················· | ···················· | ···················· | ···················· |
| K | 3252 | hCoV-19/Russia/OMS-ORINFI-6331S/2020\|EPI_ISL_56977 | ···················· | ···················· | ···················· | ···················· | ···················· | ···················· |
| K | 3253 | hCoV-19/Russia/OMS-ORINFI-9033S/2020\|EPI_ISL_56977 | ···················· | ···················· | ···················· | ···················· | ···················· | ···················· |
| K | 3254 | hCoV-19/Russia/OMS-ORINFI-1157S/2020\|EPI_ISL_56977 | ···················· | ···················· | ···················· | ···················· | ···················· | ···················· |
| K | 3255 | hCoV-19/Russia/OMS-ORINFI-2149S/2020\|EPI_ISL_56977 | ···················· | ···················· | ···················· | ···················· | ···················· | ···················· |
| K | 3256 | hCoV-19/Russia/OMS-ORINFI-2911S/2020\|EPI_ISL_56977 | ···················· | ···················· | ···················· | ···················· | ···················· | ···················· |
| K | 3257 | hCoV-19/Russia/OMS-ORINFI-3012S/2020\|EPI_ISL_56977 | ···················· | ···················· | ···················· | ···················· | ···················· | ···················· |
| K | 3258 | hCoV-19/Russia/OMS-ORINFI-4822S/2020\|EPI_ISL_56980 | ···················· | ···················· | ···················· | ···················· | ···················· | ···················· |
| K | 3259 | hCoV-19/Russia/OMS-ORINFI-8941S/2020\|EPI_ISL_56980 | ···················· | ···················· | ···················· | ···················· | ···················· | ···················· |
| K | 3260 | hCoV-19/Russia/OMS-ORINFI-61S/2020\|EPI_ISL_569802\| | ···················· | ···················· | ···················· | ···················· | ···················· | ···················· |
| K | 3261 | hCoV-19/Russia/OMS-ORINFI-5912S/2020\|EPI_ISL_56980 | ···················· | ···················· | ···················· | ···················· | ···················· | ···················· |
| K | 3262 | hCoV-19/Russia/OMS-ORINFI-4882S/2020\|EPI_ISL_56980 | ···················· | ···················· | ···················· | ···················· | ···················· | ···················· |
| K | 3263 | hCoV-19/Russia/OMS-ORINFI-3019S/2020\|EPI_ISL_56980 | ···················· | ···················· | ···················· | ···················· | ···················· | ···················· |
| K | 3264 | hCoV-19/Russia/OMS-ORINFI-4827S/2020\|EPI_ISL_56980 | ···················· | ···················· | ···················· | ···················· | ···················· | ···················· |
| K | 3265 | hCoV-19/Russia/OMS-ORINFI-6240S/2020\|EPI_ISL_56980 | ···················· | ···················· | ···················· | ···················· | ···················· | ···················· |

| | | | | | | | |
|---|---|---|---|---|---|---|---|
| K | 3266 hCoV-19/Russia/OMS-ORINFI-8942S/2020\|EPI_ISL_56980 | ···················· | ···················· | ···················· | ···················· | ···················· | ···················· |
| K | 3267 hCoV-19/Russia/OMS-ORINFI-70S/2020\|EPI_ISL_569809\| | ···················· | ···················· | ···················· | ···················· | ···················· | ···················· |
| K | 3268 hCoV-19/Russia/OMS-ORINFI-56S-2/2020\|EPI_ISL_56981 | ···················· | ···················· | ···················· | ···················· | ···················· | ···················· |
| K | 3269 hCoV-19/Russia/OMS-ORINFI-4877S/2020\|EPI_ISL_56981 | ···················· | ···················· | ···················· | ···················· | ···················· | ···················· |
| K | 3270 hCoV-19/Russia/OMS-ORINFI-66S/2020\|EPI_ISL_569812\| | ···················· | ···················· | ···················· | ···················· | ···················· | ···················· |
| K | 3271 hCoV-19/USA/WA-UW-22505/2020\|EPI_ISL_570700\|2020-0 | ···················· | ·A·················· | ···················· | ···················· | ···················· | ···················· |
| K | 3272 hCoV-19/USA/WA-UW-22514/2020\|EPI_ISL_570701\|2020-0 | ···················· | ···················· | ···················· | ···················· | ···················· | ···················· |
| K | 3273 hCoV-19/USA/WA-UW-22517/2020\|EPI_ISL_570702\|2020-0 | ···················· | ···················· | ···················· | ···················· | ···················· | ···················· |
| K | 3274 hCoV-19/USA/WA-UW-22720/2020\|EPI_ISL_570703\|2020-0 | ···················· | ···················· | ···················· | ···················· | ···················· | ···················· |
| K | 3275 hCoV-19/USA/WA-UW-22735/2020\|EPI_ISL_570704\|2020-0 | ···················· | ·A·················· | ···················· | ···················· | ···················· | ···················· |
| K | 3276 hCoV-19/USA/WA-UW-22746/2020\|EPI_ISL_570705\|2020-0 | ···················· | ···················· | ···················· | ···················· | ···················· | ···················· |
| K | 3277 hCoV-19/USA/WA-UW-22747/2020\|EPI_ISL_570706\|2020-0 | ···················· | ···················· | ···················· | ···················· | ···················· | ···················· |
| K | 3278 hCoV-19/USA/WA-UW-22748/2020\|EPI_ISL_570707\|2020-0 | ···················· | ···················· | ···················· | ···················· | ···················· | ···················· |
| K | 3279 hCoV-19/USA/WA-UW-22751/2020\|EPI_ISL_570708\|2020-0 | ···················· | ·A·················· | ···················· | ···················· | ···················· | ···················· |
| K | 3280 hCoV-19/USA/WA-UW-22752/2020\|EPI_ISL_570709\|2020-0 | ···················· | ·A·················· | ···················· | ···················· | ···················· | ···················· |
| K | 3281 hCoV-19/USA/WA-UW-22753/2020\|EPI_ISL_570710\|2020-0 | ···················· | ···················· | ···················· | ···················· | ···················· | ···················· |
| K | 3282 hCoV-19/USA/WA-UW-22756/2020\|EPI_ISL_570711\|2020-0 | ···················· | ·A·················· | ···················· | ···················· | ···················· | ···················· |
| K | 3283 hCoV-19/USA/WA-UW-22762/2020\|EPI_ISL_570714\|2020-0 | ···················· | ···················· | ···················· | ···················· | ···················· | ···················· |
| K | 3284 hCoV-19/USA/WA-UW-22769/2020\|EPI_ISL_570715\|2020-0 | ···················· | ·A·················· | ···················· | ···················· | ···················· | ···················· |
| K | 3285 hCoV-19/USA/WA-UW-22774/2020\|EPI_ISL_570716\|2020-0 | ···················· | ···················· | ···················· | ···················· | ···················· | ···················· |
| K | 3286 hCoV-19/USA/WA-UW-22782/2020\|EPI_ISL_570717\|2020-0 | ···················· | ···················· | ···················· | ···················· | ···················· | ···················· |
| K | 3287 hCoV-19/USA/WA-UW-22784/2020\|EPI_ISL_570718\|2020-0 | ···················· | ···················· | ···················· | ···················· | ···················· | ···················· |
| K | 3288 hCoV-19/USA/WA-UW-22791/2020\|EPI_ISL_570719\|2020-0 | ···················· | ···················· | ···················· | ···················· | ···················· | ···················· |
| K | 3289 hCoV-19/USA/WA-UW-22804/2020\|EPI_ISL_570720\|2020-0 | ···················· | ···················· | ···················· | ···················· | ···················· | ···················· |
| K | 3290 hCoV-19/USA/WA-UW-22827/2020\|EPI_ISL_570721\|2020-0 | ···················· | ···················· | ···················· | ···················· | ···················· | ···················· |
| K | 3291 hCoV-19/USA/WA-UW-22835/2020\|EPI_ISL_570722\|2020-0 | ···················· | ···················· | ···················· | ···················· | ···················· | ···················· |
| K | 3292 hCoV-19/USA/WA-UW-22839/2020\|EPI_ISL_570723\|2020-0 | ···················· | ·A·················· | ···················· | ···················· | ···················· | ···················· |
| K | 3293 hCoV-19/USA/WA-UW-22848/2020\|EPI_ISL_570724\|2020-0 | ···················· | ···················· | ···················· | ···················· | ···················· | ···················· |
| K | 3294 hCoV-19/USA/WA-UW-22851/2020\|EPI_ISL_570725\|2020-0 | ···················· | ···················· | ···················· | ···················· | ···················· | ···················· |
| K | 3295 hCoV-19/USA/WA-UW-22854/2020\|EPI_ISL_570726\|2020-0 | ···················· | ···················· | ···················· | ···················· | ···················· | ···················· |
| K | 3296 hCoV-19/USA/WA-UW-22859/2020\|EPI_ISL_570727\|2020-0 | ···················· | ···················· | ···················· | ···················· | ···················· | ···················· |
| K | 3297 hCoV-19/USA/WA-UW-22867/2020\|EPI_ISL_570728\|2020-0 | ···················· | ···················· | ···················· | ···················· | ···················· | ···················· |
| K | 3298 hCoV-19/USA/WA-UW-22885/2020\|EPI_ISL_570729\|2020-0 | ···················· | ·A·················· | ···················· | ···················· | ···················· | ···················· |
| K | 3299 hCoV-19/USA/WA-UW-22892/2020\|EPI_ISL_570730\|2020-0 | ···················· | ·A·················· | ···················· | ···················· | ···················· | ···················· |
| K | 3300 hCoV-19/USA/WA-UW-22899/2020\|EPI_ISL_570731\|2020-0 | ···················· | ···················· | ···················· | ···················· | ···················· | ···················· |
| K | 3301 hCoV-19/USA/WA-UW-22900/2020\|EPI_ISL_570732\|2020-0 | ···················· | ···················· | ···················· | ···················· | ···················· | ···················· |
| K | 3302 hCoV-19/USA/WA-UW-22903/2020\|EPI_ISL_570733\|2020-0 | ···················· | ···················· | ···················· | ···················· | ···················· | ···················· |
| K | 3303 hCoV-19/USA/WA-UW-22920/2020\|EPI_ISL_570734\|2020-0 | ···················· | ···················· | ···················· | ···················· | ···················· | ···················· |
| K | 3304 hCoV-19/USA/WA-UW-22922/2020\|EPI_ISL_570735\|2020-0 | ···················· | ···················· | ···················· | ···················· | ···················· | ···················· |
| K | 3305 hCoV-19/USA/WA-UW-22948/2020\|EPI_ISL_570736\|2020-0 | ···················· | ·A·················· | ···················· | ···················· | ···················· | ···················· |
| K | 3306 hCoV-19/USA/WA-UW-22951/2020\|EPI_ISL_570737\|2020-0 | ···················· | ·A·················· | ···················· | ···················· | ···················· | ···················· |
| K | 3307 hCoV-19/USA/WA-UW-22952/2020\|EPI_ISL_570738\|2020-0 | ···················· | ···················· | ···················· | ···················· | ···················· | ···················· |
| K | 3308 hCoV-19/USA/WA-UW-22963/2020\|EPI_ISL_570739\|2020-0 | ···················· | ···················· | ···················· | ···················· | ···················· | ···················· |
| K | 3309 hCoV-19/USA/WA-UW-22972/2020\|EPI_ISL_570740\|2020-0 | ···················· | ·A·················· | ···················· | ···················· | ···················· | ···················· |
| K | 3310 hCoV-19/USA/WA-UW-22978/2020\|EPI_ISL_570741\|2020-0 | ···················· | ·A·················· | ···················· | ···················· | ···················· | ···················· |
| K | 3311 hCoV-19/USA/WA-UW-22981/2020\|EPI_ISL_570742\|2020-0 | ···················· | ·A·················· | ···················· | ···················· | ···················· | ···················· |
| K | 3312 hCoV-19/USA/WA-UW-22990/2020\|EPI_ISL_570743\|2020-0 | ···················· | ···················· | ···················· | ···················· | ···················· | ···················· |
| K | 3313 hCoV-19/USA/WA-UW-22993/2020\|EPI_ISL_570744\|2020-0 | ···················· | ···················· | ···················· | ···················· | ···················· | ···················· |
| K | 3314 hCoV-19/USA/WA-UW-22994/2020\|EPI_ISL_570745\|2020-0 | ···················· | ···················· | ···················· | ···················· | ···················· | ···················· |
| K | 3315 hCoV-19/USA/WA-UW-22998/2020\|EPI_ISL_570746\|2020-0 | ···················· | ···················· | ···················· | ···················· | ···················· | ···················· |
| K | 3316 hCoV-19/USA/WA-UW-23004/2020\|EPI_ISL_570747\|2020-0 | ···················· | ···················· | ···················· | ···················· | ···················· | ···················· |
| K | 3317 hCoV-19/USA/WA-UW-23124/2020\|EPI_ISL_570748\|2020-0 | ···················· | ···················· | ···················· | ···················· | ···················· | ···················· |
| K | 3318 hCoV-19/USA/WA-UW-23126/2020\|EPI_ISL_570749\|2020-0 | ···················· | ···················· | ···················· | ···················· | ···················· | ···················· |
| K | 3319 hCoV-19/USA/WA-UW-23130/2020\|EPI_ISL_570750\|2020-0 | ···················· | ·A·················· | ···················· | ···················· | ···················· | ···················· |
| K | 3320 hCoV-19/USA/WA-UW-23152/2020\|EPI_ISL_570751\|2020-0 | ···················· | ·A·················· | ···················· | ···················· | ···················· | ···················· |
| K | 3321 hCoV-19/USA/WA-UW-23164/2020\|EPI_ISL_570752\|2020-0 | ···················· | ·A·················· | ···················· | ···················· | ···················· | ···················· |
| K | 3322 hCoV-19/USA/WA-UW-23184/2020\|EPI_ISL_570753\|2020-0 | ···················· | ···················· | ···················· | ···················· | ···················· | ···················· |
| K | 3323 hCoV-19/USA/WA-UW-23193/2020\|EPI_ISL_570754\|2020-0 | ···················· | ···················· | ···················· | ···················· | ···················· | ···················· |
| K | 3324 hCoV-19/USA/WA-UW-23196/2020\|EPI_ISL_570755\|2020-0 | ···················· | ·A·················· | ···················· | ···················· | ···················· | ···················· |
| K | 3325 hCoV-19/USA/WA-UW-23200/2020\|EPI_ISL_570756\|2020-0 | ···················· | ···················· | ···················· | ···················· | ···················· | ···················· |
| K | 3326 hCoV-19/USA/WA-UW-23206/2020\|EPI_ISL_570757\|2020-0 | ···················· | ···················· | ···················· | ···················· | ···················· | ···················· |
| K | 3327 hCoV-19/USA/WA-UW-23213/2020\|EPI_ISL_570758\|2020-0 | ···················· | ·A·················· | ···················· | ···················· | ···················· | ···················· |
| K | 3328 hCoV-19/USA/WA-UW-23483/2020\|EPI_ISL_570759\|2020-0 | ···················· | ·A·················· | ···················· | ···················· | ···················· | ···················· |
| K | 3329 hCoV-19/Gambia/3683/2020\|EPI_ISL_561000\|2020-07-30 | ···················· | ···················· | ···················· | ···················· | ···················· | ···················· |
| K | 3330 hCoV-19/Gambia/3703/2020\|EPI_ISL_561001\|2020-07-30 | ···················· | ···················· | ···················· | ···················· | ···················· | ···················· |
| K | 3331 hCoV-19/Gambia/3724/2020\|EPI_ISL_561003\|2020-07-30 | ···················· | ···················· | ···················· | ···················· | ···················· | ···················· |
| K | 3332 hCoV-19/Gambia/3725/2020\|EPI_ISL_561004\|2020-07-30 | ···················· | ···················· | ···················· | ···················· | ···················· | ···················· |
| K | 3333 hCoV-19/Gambia/3729/2020\|EPI_ISL_561006\|2020-07-30 | ···················· | ···················· | ···················· | ···················· | ···················· | ···················· |
| K | 3334 hCoV-19/Gambia/3743/2020\|EPI_ISL_561011\|2020-07-30 | ···················· | ···················· | ···················· | ···················· | ···················· | ···················· |
| K | 3335 hCoV-19/Gambia/3749/2020\|EPI_ISL_561012\|2020-07-30 | ···················· | ···················· | ···················· | ···················· | ···················· | ···················· |
| K | 3336 hCoV-19/Australia/VIC12839/2020\|EPI_ISL_563600\|202 | ···················· | ···················· | ···················· | ···················· | ···················· | ···················· |
| K | 3337 hCoV-19/Australia/VIC12851/2020\|EPI_ISL_563601\|202 | ···················· | ···················· | ···················· | ···················· | ···················· | ···················· |
| K | 3338 hCoV-19/Australia/VIC12870/2020\|EPI_ISL_563602\|202 | ···················· | ···················· | ···················· | ···················· | ···················· | ···················· |
| K | 3339 hCoV-19/Australia/VIC12873/2020\|EPI_ISL_563603\|202 | ···················· | ···················· | ···················· | ···················· | ···················· | ···················· |
| K | 3341 hCoV-19/Australia/VIC12913/2020\|EPI_ISL_563605\|202 | ···················· | ···················· | ···················· | ···················· | ···················· | ···················· |
| K | 3342 hCoV-19/Australia/VIC12922/2020\|EPI_ISL_563607\|202 | ···················· | ···················· | ···················· | ···················· | ···················· | ···················· |
| K | 3343 hCoV-19/Australia/VIC12923/2020\|EPI_ISL_563608\|202 | ···················· | ···················· | ···················· | ···················· | ···················· | ···················· |
| K | 3345 hCoV-19/Australia/VIC12949/2020\|EPI_ISL_563610\|202 | ···················· | ···················· | ···················· | ···················· | ···················· | ···················· |
| K | 3346 hCoV-19/Australia/VIC12955/2020\|EPI_ISL_563611\|202 | ···················· | ···················· | ···················· | ···················· | ···················· | ···················· |
| K | 3347 hCoV-19/Australia/VIC12960/2020\|EPI_ISL_563612\|202 | ···················· | ···················· | ···················· | ···················· | ···················· | NNN················· |

| | | | | | | | | |
|---|---|---|---|---|---|---|---|---|
| K | 3348 hCoV-19/Australia/VIC12966/2020\|EPI_ISL_563613\|202 | ···················· | ···················· | ····················· | ···················· | ···················· | ···················· | |
| K | 3349 hCoV-19/Australia/VIC12975/2020\|EPI_ISL_563614\|202 | ···················· | ···················· | ····················· | ···················· | ···················· | ···················· | |
| K | 3350 hCoV-19/Australia/VIC12977/2020\|EPI_ISL_563615\|202 | ···················· | ···················· | ····················· | ···················· | ···················· | ···················· | |
| K | 3351 hCoV-19/Australia/VIC13066/2020\|EPI_ISL_563616\|202 | ···················· | ···················· | ····················· | ···················· | ···················· | ···················· | |
| K | 3352 hCoV-19/Australia/VIC13082/2020\|EPI_ISL_563617\|202 | ···················· | ···················· | ····················· | ···················· | ···················· | ···················· | |
| K | 3353 hCoV-19/Australia/VIC13087/2020\|EPI_ISL_563618\|202 | ···················· | ···················· | ····················· | ···················· | ···················· | ···················· | |
| K | 3354 hCoV-19/Australia/VIC13093/2020\|EPI_ISL_563619\|202 | ···················· | ···················· | ····················· | ···················· | ···················· | ···················· | |
| K | 3355 hCoV-19/Australia/VIC13108/2020\|EPI_ISL_563620\|202 | ···················· | ···················· | ····················· | ···················· | ···················· | ···················· | |
| K | 3356 hCoV-19/Australia/VIC13112/2020\|EPI_ISL_563621\|202 | ···················· | ···················· | ····················· | ···················· | ···················· | ···················· | |
| K | 3357 hCoV-19/Australia/VIC13122/2020\|EPI_ISL_563622\|202 | ···················· | ···················· | ····················· | ···················· | ···················· | ···················· | |
| K | 3358 hCoV-19/Australia/VIC13138/2020\|EPI_ISL_563623\|202 | ···················· | ···················· | ····················· | ···················· | ···················· | ···················· | |
| K | 3359 hCoV-19/Australia/VIC13140/2020\|EPI_ISL_563624\|202 | ···················· | ···················· | ····················· | ···················· | ···················· | ···················· | |
| K | 3360 hCoV-19/Australia/VIC13146/2020\|EPI_ISL_563625\|202 | ···················· | ···················· | ····················· | ···················· | ···················· | ···················· | |
| K | 3361 hCoV-19/Australia/VIC13153/2020\|EPI_ISL_563626\|202 | ···················· | ···················· | ····················· | ···················· | ···················· | ···················· | |
| K | 3362 hCoV-19/Australia/VIC13156/2020\|EPI_ISL_563627\|202 | ···················· | ···················· | ····················· | ···················· | ···················· | ···················· | |
| K | 3363 hCoV-19/Australia/VIC13178/2020\|EPI_ISL_563628\|202 | ···················· | ···················· | ····················· | ···················· | ···················· | ···················· | |
| K | 3364 hCoV-19/Australia/VIC13180/2020\|EPI_ISL_563629\|202 | ···················· | ···················· | ····················· | ···················· | ···················· | ···················· | |
| K | 3365 hCoV-19/Australia/VIC13181/2020\|EPI_ISL_563630\|202 | ···················· | ···················· | ····················· | ···················· | ···················· | ···················· | |
| K | 3366 hCoV-19/Australia/VIC13182/2020\|EPI_ISL_563631\|202 | ···················· | ···················· | ····················· | ···················· | ···················· | ···················· | |
| K | 3367 hCoV-19/Australia/VIC13183/2020\|EPI_ISL_563632\|202 | ···················· | ···················· | ····················· | ···················· | ···················· | ···················· | |
| K | 3368 hCoV-19/Australia/VIC13184/2020\|EPI_ISL_563633\|202 | ···················· | ···················· | ····················· | ···················· | ···················· | ···················· | |
| K | 3369 hCoV-19/Australia/VIC13186/2020\|EPI_ISL_563634\|202 | ···················· | ···················· | ····················· | ···················· | ···················· | ···················· | |
| K | 3370 hCoV-19/Australia/VIC13188/2020\|EPI_ISL_563635\|202 | ···················· | ···················· | ····················· | ···················· | ···················· | ···················· | |
| K | 3371 hCoV-19/Australia/VIC13189/2020\|EPI_ISL_563636\|202 | ···················· | ···················· | ····················· | ···················· | ···················· | ···················· | |
| K | 3372 hCoV-19/Australia/VIC13195/2020\|EPI_ISL_563637\|202 | ···················· | ···················· | ····················· | ···················· | ···················· | ···················· | |
| K | 3373 hCoV-19/Australia/VIC13196/2020\|EPI_ISL_563638\|202 | ···················· | ···················· | ····················· | ···················· | ···················· | ···················· | |
| K | 3374 hCoV-19/Australia/VIC13199/2020\|EPI_ISL_563639\|202 | ···················· | ···················· | ····················· | ···················· | ···················· | ···················· | |
| K | 3375 hCoV-19/Australia/VIC13205/2020\|EPI_ISL_563640\|202 | ···················· | ···················· | ····················· | ···················· | ···················· | ···················· | |
| K | 3376 hCoV-19/Australia/VIC13210/2020\|EPI_ISL_563641\|202 | ···················· | ···················· | ····················· | ···················· | ···················· | ···················· | |
| K | 3377 hCoV-19/Australia/VIC13223/2020\|EPI_ISL_563642\|202 | ···················· | ···················· | ····················· | ···················· | ···················· | ···················· | |
| K | 3378 hCoV-19/Australia/VIC13227/2020\|EPI_ISL_563643\|202 | ···················· | ···················· | ····················· | ···················· | ···················· | ···················· | |
| K | 3379 hCoV-19/Australia/VIC13236/2020\|EPI_ISL_563644\|202 | ···················· | ···················· | ····················· | ···················· | ···················· | ···················· | |
| K | 3380 hCoV-19/Australia/VIC13238/2020\|EPI_ISL_563645\|202 | ···················· | ···················· | ····················· | ···················· | ···················· | ···················· | |
| K | 3381 hCoV-19/Australia/VIC13245/2020\|EPI_ISL_563646\|202 | ···················· | ···················· | ····················· | ···················· | ···················· | ···················· | |
| K | 3382 hCoV-19/Australia/VIC13268/2020\|EPI_ISL_563647\|202 | ···················· | ···················· | ····················· | ···················· | ···················· | ···················· | |
| K | 3383 hCoV-19/Australia/VIC13271/2020\|EPI_ISL_563648\|202 | ···················· | ···················· | ····················· | ···················· | ···················· | ···················· | |
| K | 3384 hCoV-19/Australia/VIC13279/2020\|EPI_ISL_563649\|202 | ···················· | ···················· | ····················· | ···················· | ···················· | ···················· | |
| K | 3385 hCoV-19/Australia/VIC13282/2020\|EPI_ISL_563650\|202 | ···················· | ···················· | ····················· | ···················· | ···················· | ···················· | |
| K | 3386 hCoV-19/Australia/VIC13283/2020\|EPI_ISL_563651\|202 | ···················· | ···················· | ····················· | ···················· | ···················· | ···················· | |
| K | 3387 hCoV-19/Australia/VIC13284/2020\|EPI_ISL_563652\|202 | ···················· | ···················· | ····················· | ···················· | ···················· | ···················· | |
| K | 3388 hCoV-19/Australia/VIC13286/2020\|EPI_ISL_563653\|202 | ···················· | ···················· | ····················· | ···················· | ···················· | ···················· | |
| K | 3389 hCoV-19/Australia/VIC13293/2020\|EPI_ISL_563654\|202 | ···················· | ···················· | ····················· | ···················· | ···················· | ···················· | |
| K | 3390 hCoV-19/Australia/VIC13295/2020\|EPI_ISL_563655\|202 | ···················· | ···················· | ····················· | ···················· | ···················· | ···················· | |
| K | 3391 hCoV-19/Australia/VIC13296/2020\|EPI_ISL_563656\|202 | ···················· | ···················· | ····················· | ···················· | ···················· | ···················· | |
| K | 3392 hCoV-19/Australia/VIC13297/2020\|EPI_ISL_563657\|202 | ···················· | ···················· | ····················· | ···················· | ···················· | ···················· | |
| K | 3393 hCoV-19/Australia/VIC13298/2020\|EPI_ISL_563658\|202 | ···················· | ···················· | ····················· | ···················· | ···················· | ···················· | |
| K | 3394 hCoV-19/Australia/VIC13300/2020\|EPI_ISL_563659\|202 | ···················· | ···················· | ····················· | ···················· | ···················· | ···················· | |
| K | 3395 hCoV-19/Australia/VIC13301/2020\|EPI_ISL_563660\|202 | ···················· | ···················· | ····················· | ···················· | ···················· | ···················· | |
| K | 3396 hCoV-19/Australia/VIC13303/2020\|EPI_ISL_563661\|202 | ···················· | ···················· | ····················· | ···················· | ···················· | ···················· | |
| K | 3397 hCoV-19/Australia/VIC13306/2020\|EPI_ISL_563662\|202 | ···················· | ···················· | ····················· | ···················· | ···················· | ···················· | |
| K | 3398 hCoV-19/Australia/VIC13307/2020\|EPI_ISL_563663\|202 | ···················· | ···················· | ····················· | ···················· | ···················· | NNN················· | 1 |
| K | 3399 hCoV-19/Australia/VIC13312/2020\|EPI_ISL_563664\|202 | ···················· | ···················· | ····················· | ···················· | ···················· | ···················· | |
| K | 3400 hCoV-19/Australia/VIC13313/2020\|EPI_ISL_563665\|202 | ···················· | ···G················ | ····················· | ···················· | ···················· | ···················· | |
| K | 3401 hCoV-19/Australia/VIC13314/2020\|EPI_ISL_563666\|202 | ···················· | ···················· | ····················· | ···················· | ···················· | ···················· | |
| K | 3402 hCoV-19/Australia/VIC13316/2020\|EPI_ISL_563667\|202 | ···················· | ···················· | ····················· | ···················· | ···················· | ···················· | |
| K | 3403 hCoV-19/Australia/VIC13317/2020\|EPI_ISL_563668\|202 | ···················· | ···················· | ····················· | ···················· | ···················· | ···················· | |
| K | 3404 hCoV-19/Australia/VIC13318/2020\|EPI_ISL_563669\|202 | ···················· | ···················· | ····················· | ···················· | ···················· | ···················· | |
| K | 3405 hCoV-19/Australia/VIC13319/2020\|EPI_ISL_563670\|202 | ···················· | ···················· | ····················· | ···················· | ···················· | ···················· | |
| K | 3407 hCoV-19/Australia/VIC13321/2020\|EPI_ISL_563672\|202 | ···················· | ···················· | ····················· | ···················· | ···················· | ···················· | |
| K | 3408 hCoV-19/Australia/VIC13325/2020\|EPI_ISL_563673\|202 | ···················· | ···················· | ····················· | ···················· | ···················· | ···················· | |
| K | 3409 hCoV-19/Australia/VIC13326/2020\|EPI_ISL_563674\|202 | ···················· | ···················· | ····················· | ···················· | ···················· | ···················· | |
| K | 3410 hCoV-19/Australia/VIC8254/2020\|EPI_ISL_564920\|2020 | ···················· | ···················· | ····················· | ···················· | ···················· | ···················· | |
| K | 3411 hCoV-19/Australia/VIC8255/2020\|EPI_ISL_564921\|2020 | ···················· | ···················· | ····················· | ···················· | ···················· | ···················· | |
| K | 3412 hCoV-19/Australia/VIC8261/2020\|EPI_ISL_564922\|2020 | ···················· | ···················· | ····················· | ···················· | ···················· | ···················· | |
| K | 3413 hCoV-19/Australia/VIC8266/2020\|EPI_ISL_564923\|2020 | ···················· | ···················· | ····················· | ···················· | ···················· | ···················· | |
| K | 3414 hCoV-19/Australia/VIC8270/2020\|EPI_ISL_564924\|2020 | ···················· | ···················· | ····················· | ···················· | ···················· | ···················· | |
| K | 3415 hCoV-19/Australia/VIC8278/2020\|EPI_ISL_564925\|2020 | ···················· | ···················· | ····················· | ···················· | ···················· | ···················· | |
| K | 3416 hCoV-19/Australia/VIC8280/2020\|EPI_ISL_564926\|2020 | ···················· | ···················· | ····················· | ···················· | ···················· | ···················· | |
| K | 3417 hCoV-19/Australia/VIC8281/2020\|EPI_ISL_564927\|2020 | ···················· | ···················· | ····················· | ···················· | ···················· | ···················· | |
| K | 3418 hCoV-19/Australia/VIC8293/2020\|EPI_ISL_564928\|2020 | ···················· | ···················· | ····················· | ···················· | ···················· | ···················· | |
| K | 3419 hCoV-19/Australia/VIC8297/2020\|EPI_ISL_564929\|2020 | ···················· | ···················· | ····················· | ···················· | ···················· | ···················· | |
| K | 3420 hCoV-19/Australia/VIC8303/2020\|EPI_ISL_564930\|2020 | ···················· | ···················· | ····················· | ···················· | ···················· | ···················· | |
| K | 3421 hCoV-19/Australia/VIC8304/2020\|EPI_ISL_564931\|2020 | ···················· | ···················· | ····················· | ···················· | ···················· | ···················· | |
| K | 3422 hCoV-19/Australia/VIC8305/2020\|EPI_ISL_564932\|2020 | ···················· | ···················· | ····················· | ···················· | ···················· | ···················· | |
| K | 3423 hCoV-19/Australia/VIC8306/2020\|EPI_ISL_564933\|2020 | ···················· | ···················· | ····················· | ···················· | ···················· | ···················· | |
| K | 3424 hCoV-19/Australia/VIC8309/2020\|EPI_ISL_564934\|2020 | ···················· | ···················· | ····················· | ···················· | ···················· | ···················· | |
| K | 3425 hCoV-19/Australia/VIC8311/2020\|EPI_ISL_564935\|2020 | ···················· | ···················· | ····················· | ···················· | ···················· | ···················· | |
| K | 3426 hCoV-19/Australia/VIC8314/2020\|EPI_ISL_564936\|2020 | ···················· | ···················· | ····················· | ···················· | ···················· | ···················· | |
| K | 3427 hCoV-19/Australia/VIC8323/2020\|EPI_ISL_564939\|2020 | ···················· | ···················· | ····················· | ···················· | ···················· | ···················· | |
| K | 3428 hCoV-19/Australia/VIC8324/2020\|EPI_ISL_564940\|2020 | ···················· | ···················· | ····················· | ···················· | ···················· | ···················· | |

| | | | | | | | |
|---|---|---|---|---|---|---|---|
| K | 3429 hCoV-19/Australia/VIC8325/2020\|EPI_ISL_564941\|2020 | ···················· | ···················· | ···················· | ···················· | ···················· | ···················· |
| K | 3430 hCoV-19/Australia/VIC8327/2020\|EPI_ISL_564942\|2020 | ···················· | ···················· | ···················· | ···················· | ···················· | ···················· |
| K | 3431 hCoV-19/Australia/VIC8328/2020\|EPI_ISL_564943\|2020 | ···················· | ···················· | ···················· | ···················· | ···················· | ···················· |
| K | 3432 hCoV-19/Australia/VIC8330/2020\|EPI_ISL_564944\|2020 | ···················· | ···················· | ···················· | ···················· | ···················· | ···················· |
| K | 3433 hCoV-19/Australia/VIC8331/2020\|EPI_ISL_564945\|2020 | ···················· | ···················· | ···················· | ···················· | ···················· | ···················· |
| K | 3434 hCoV-19/Australia/VIC8333/2020\|EPI_ISL_564946\|2020 | ···················· | ···················· | ···················· | ···················· | ···················· | ···················· |
| K | 3435 hCoV-19/Australia/VIC8334/2020\|EPI_ISL_564947\|2020 | ···················· | ···················· | ···················· | ···················· | ···················· | ···················· |
| K | 3436 hCoV-19/Australia/VIC8335/2020\|EPI_ISL_564948\|2020 | ···················· | ···················· | ···················· | ···················· | ···················· | ···················· |
| K | 3437 hCoV-19/Australia/VIC8336/2020\|EPI_ISL_564949\|2020 | ···················· | ···················· | ···················· | ···················· | ···················· | ···················· |
| K | 3438 hCoV-19/Australia/VIC8337/2020\|EPI_ISL_564950\|2020 | ···················· | ···················· | ···················· | ···················· | ···················· | ···················· |
| K | 3439 hCoV-19/Australia/VIC8339/2020\|EPI_ISL_564951\|2020 | ···················· | ···················· | ···················· | ···················· | ···················· | ···················· |
| K | 3440 hCoV-19/Australia/VIC8340/2020\|EPI_ISL_564952\|2020 | ···················· | ···················· | ···················· | ···················· | ···················· | ···················· |
| K | 3441 hCoV-19/Australia/VIC8341/2020\|EPI_ISL_564953\|2020 | ···················· | ···················· | ···················· | ···················· | ···················· | ···················· |
| K | 3442 hCoV-19/Australia/VIC8342/2020\|EPI_ISL_564954\|2020 | ···················· | ···················· | ···················· | ···················· | ···················· | ···················· |
| K | 3443 hCoV-19/Australia/VIC8343/2020\|EPI_ISL_564955\|2020 | ···················· | ···················· | ···················· | ···················· | ···················· | ···················· |
| K | 3444 hCoV-19/Australia/VIC8345/2020\|EPI_ISL_564956\|2020 | ···················· | ···················· | ···················· | ···················· | ···················· | ···················· |
| K | 3445 hCoV-19/Australia/VIC8346/2020\|EPI_ISL_564957\|2020 | ···················· | ···················· | ···················· | ···················· | ···················· | ···················· |
| K | 3446 hCoV-19/Australia/VIC8348/2020\|EPI_ISL_564958\|2020 | ···················· | ···················· | ···················· | ···················· | ···················· | ···················· |
| K | 3447 hCoV-19/Australia/VIC8349/2020\|EPI_ISL_564959\|2020 | ···················· | ···················· | ···················· | ···················· | ···················· | ···················· |
| K | 3448 hCoV-19/Australia/VIC8350/2020\|EPI_ISL_564960\|2020 | ···················· | ···················· | ···················· | ···················· | ···················· | ···················· |
| K | 3449 hCoV-19/Australia/VIC8352/2020\|EPI_ISL_564961\|2020 | ···················· | ···················· | ···················· | ···················· | ···················· | ···················· |
| K | 3450 hCoV-19/Australia/VIC8353/2020\|EPI_ISL_564962\|2020 | ···················· | ···················· | ···················· | ···················· | ···················· | ···················· |
| K | 3451 hCoV-19/Australia/VIC8354/2020\|EPI_ISL_564963\|2020 | ···················· | ···················· | ···················· | ···················· | ···················· | ···················· |
| K | 3452 hCoV-19/Australia/VIC8362/2020\|EPI_ISL_564964\|2020 | ···················· | ···················· | ···················· | ···················· | ···················· | ···················· |
| K | 3453 hCoV-19/Australia/VIC8363/2020\|EPI_ISL_564965\|2020 | ···················· | ···················· | ···················· | ···················· | ···················· | ···················· |
| K | 3454 hCoV-19/Australia/VIC8364/2020\|EPI_ISL_564966\|2020 | ···················· | ···················· | ···················· | ···················· | ···················· | ···················· |
| K | 3455 hCoV-19/Australia/VIC8366/2020\|EPI_ISL_564967\|2020 | ···················· | ···················· | ···················· | ···················· | ···················· | ···················· |
| K | 3456 hCoV-19/Australia/VIC8367/2020\|EPI_ISL_564968\|2020 | ···················· | ···················· | ···················· | ···················· | ···················· | ···················· |
| K | 3457 hCoV-19/Australia/VIC8368/2020\|EPI_ISL_564969\|2020 | ···················· | ···················· | ···················· | ···················· | ···················· | ···················· |
| K | 3458 hCoV-19/Australia/VIC8370/2020\|EPI_ISL_564970\|2020 | ········T··········· | ···················· | ···················· | ···················· | ···················· | ···················· |
| K | 3459 hCoV-19/Australia/VIC8374/2020\|EPI_ISL_564971\|2020 | ···················· | ···················· | ···················· | ···················· | ···················· | ···················· |
| K | 3460 hCoV-19/Australia/VIC8375/2020\|EPI_ISL_564972\|2020 | ···················· | ···················· | ···················· | ···················· | ···················· | ···················· |
| K | 3461 hCoV-19/Australia/VIC8379/2020\|EPI_ISL_564973\|2020 | ···················· | ···················· | ···················· | ···················· | ···················· | ···················· |
| K | 3462 hCoV-19/Australia/VIC8382/2020\|EPI_ISL_564974\|2020 | ···················· | ···················· | ···················· | ···················· | ···················· | ···················· |
| K | 3463 hCoV-19/Australia/VIC8385/2020\|EPI_ISL_564975\|2020 | ···················· | ···················· | ···················· | ···················· | ···················· | ···················· |
| K | 3464 hCoV-19/Australia/VIC8386/2020\|EPI_ISL_564976\|2020 | ···················· | ···················· | ···················· | ···················· | ···················· | ···················· |
| K | 3465 hCoV-19/Australia/VIC8387/2020\|EPI_ISL_564977\|2020 | ···················· | ···················· | ···················· | ···················· | ···················· | ···················· |
| K | 3466 hCoV-19/Australia/VIC8389/2020\|EPI_ISL_564978\|2020 | ···················· | ···················· | ···················· | ···················· | ···················· | ···················· |
| K | 3467 hCoV-19/Australia/VIC8391/2020\|EPI_ISL_564979\|2020 | ···················· | ···················· | ···················· | ···················· | ···················· | ···················· |
| K | 3468 hCoV-19/Australia/VIC8392/2020\|EPI_ISL_564980\|2020 | ···················· | ···················· | ···················· | ···················· | ···················· | ···················· |
| K | 3469 hCoV-19/Australia/VIC8393/2020\|EPI_ISL_564981\|2020 | ···················· | ···················· | ···················· | ···················· | ···················· | ···················· |
| K | 3470 hCoV-19/Australia/VIC8394/2020\|EPI_ISL_564982\|2020 | ···················· | ···················· | ···················· | ···················· | ···················· | ···················· |
| K | 3471 hCoV-19/Australia/VIC8395/2020\|EPI_ISL_564983\|2020 | ···················· | ···················· | ···················· | ···················· | ···················· | ···················· |
| K | 3472 hCoV-19/Australia/VIC8396/2020\|EPI_ISL_564984\|2020 | ···················· | ···················· | ···················· | ···················· | ···················· | ···················· |
| K | 3473 hCoV-19/Australia/VIC8398/2020\|EPI_ISL_564985\|2020 | ···················· | ···················· | ···················· | ···················· | ···················· | ···················· |
| K | 3474 hCoV-19/Australia/VIC8400/2020\|EPI_ISL_564986\|2020 | ···················· | ···················· | ···················· | ···················· | ···················· | ···················· |
| K | 3475 hCoV-19/Australia/VIC8401/2020\|EPI_ISL_564987\|2020 | ···················· | ···················· | ···················· | ···················· | ···················· | ···················· |
| K | 3476 hCoV-19/Australia/VIC8402/2020\|EPI_ISL_564988\|2020 | ···················· | ···················· | ···················· | ···················· | ···················· | ···················· |
| K | 3477 hCoV-19/Australia/VIC8403/2020\|EPI_ISL_564989\|2020 | ···················· | ···················· | ···················· | ···················· | ···················· | ···················· |
| K | 3478 hCoV-19/Australia/VIC8405/2020\|EPI_ISL_564990\|2020 | ···················· | ···················· | ···················· | ···················· | ···················· | ···················· |
| K | 3479 hCoV-19/Australia/VIC8407/2020\|EPI_ISL_564991\|2020 | ···················· | ···················· | ···················· | ···················· | ···················· | ···················· |
| K | 3480 hCoV-19/Australia/VIC8408/2020\|EPI_ISL_564992\|2020 | ···················· | ···················· | ···················· | ···················· | ···················· | ···················· |
| K | 3481 hCoV-19/Australia/VIC8410/2020\|EPI_ISL_564993\|2020 | ···················· | ···················· | ···················· | ···················· | ···················· | ···················· |
| K | 3482 hCoV-19/Australia/VIC8412/2020\|EPI_ISL_564994\|2020 | ···················· | ···················· | ···················· | ···················· | ···················· | ···················· |
| K | 3483 hCoV-19/Australia/VIC8413/2020\|EPI_ISL_564995\|2020 | ···················· | ···················· | ···················· | ···················· | ···················· | ···················· |
| K | 3484 hCoV-19/Australia/VIC8415/2020\|EPI_ISL_564996\|2020 | ···················· | ···················· | ···················· | ···················· | ···················· | ···················· |
| K | 3485 hCoV-19/Australia/VIC8416/2020\|EPI_ISL_564997\|2020 | ···················· | ···················· | ···················· | ···················· | ···················· | ···················· |
| K | 3486 hCoV-19/Australia/VIC8417/2020\|EPI_ISL_564998\|2020 | ···················· | ···················· | ···················· | ···················· | ···················· | ···················· |
| K | 3487 hCoV-19/Australia/VIC8419/2020\|EPI_ISL_564999\|2020 | ···················· | ···················· | ···················· | ···················· | ···················· | ···················· |
| K | 3488 hCoV-19/England/QEUH-9C851B/2020\|EPI_ISL_567001\|20 | ···················· | ···················· | ···················· | ···················· | ······T············· | ···················· |
| K | 3489 hCoV-19/England/QEUH-9C8FFF/2020\|EPI_ISL_567002\|20 | ···················· | ···················· | ···················· | ···················· | ···················· | ···················· |
| K | 3490 hCoV-19/England/QEUH-9C828D/2020\|EPI_ISL_567003\|20 | ···················· | ···················· | ···················· | ···················· | ···················· | ···················· |
| K | 3491 hCoV-19/England/QEUH-9C86FA/2020\|EPI_ISL_567004\|20 | ···················· | ···················· | ···················· | ···················· | ···················· | ···················· |
| K | 3492 hCoV-19/England/QEUH-9C89D3/2020\|EPI_ISL_567005\|20 | ···················· | ···················· | ···················· | ···················· | ···················· | ···················· |
| K | 3493 hCoV-19/England/QEUH-9C8A49/2020\|EPI_ISL_567006\|20 | ···················· | ·A·················· | ···················· | ···················· | ···················· | ···A················ |
| K | 3494 hCoV-19/England/QEUH-9C83A8/2020\|EPI_ISL_567007\|20 | ···················· | ···················· | ···················· | ···················· | ···················· | ···················· |
| K | 3495 hCoV-19/England/QEUH-9C8399/2020\|EPI_ISL_567008\|20 | ···················· | ···················· | ···················· | ···················· | ···················· | ···················· |
| K | 3496 hCoV-19/Wales/QEUH-9C7F4B/2020\|EPI_ISL_567009\|2020 | ···················· | ···················· | ···················· | ···················· | ···················· | ···················· |
| K | 3497 hCoV-19/England/QEUH-9C7B1A/2020\|EPI_ISL_567010\|20 | ···················· | ···················· | ···················· | ···················· | ···················· | ···················· |
| K | 3498 hCoV-19/England/QEUH-9C7B47/2020\|EPI_ISL_567011\|20 | ···················· | ···················· | ···················· | ···················· | ···················· | ···················· |
| K | 3499 hCoV-19/England/QEUH-9C7C44/2020\|EPI_ISL_567012\|20 | ···················· | ····A··············· | ···················· | ···················· | ···················· | ···················· |
| K | 3500 hCoV-19/England/QEUH-9C8566/2020\|EPI_ISL_567013\|20 | ···················· | ···················· | ···················· | ···················· | ···················· | ···················· |
| K | 3501 hCoV-19/England/QEUH-9C8672/2020\|EPI_ISL_567014\|20 | ···················· | ···················· | ···················· | ···················· | ···················· | ···················· |
| K | 3502 hCoV-19/England/QEUH-9C7DD8/2020\|EPI_ISL_567015\|20 | ···················· | ···················· | ···················· | ···················· | ···················· | ···················· |
| K | 3503 hCoV-19/Northern Ireland/QEUH-9C85A2/2020\|EPI_ISL_ | ···················· | ···················· | ···················· | ···················· | ···················· | ···················· |
| K | 3504 hCoV-19/England/QEUH-9C8D7D/2020\|EPI_ISL_567017\|20 | ···················· | ···················· | ···················· | ···················· | ···················· | ···················· |
| K | 3505 hCoV-19/England/QEUH-9C837B/2020\|EPI_ISL_567018\|20 | ···················· | ···················· | ···················· | ···················· | ···················· | ···················· |
| K | 3506 hCoV-19/England/QEUH-9C7B83/2020\|EPI_ISL_567019\|20 | ···················· | ···················· | ···················· | ···················· | ···················· | ···················· |
| K | 3507 hCoV-19/England/QEUH-9C8548/2020\|EPI_ISL_567030\|20 | ···················· | ···················· | ···················· | ···················· | ···················· | ···················· |
| K | 3508 hCoV-19/England/QEUH-9C8162/2020\|EPI_ISL_567031\|20 | ···················· | ···················· | ···················· | ···················· | ···················· | ···················· |

```
K    3509 hCoV-19/England/QEUH-9C8593/2020|EPI_ISL_567033|20    ....................  .A..................  ....................  ....................  ....................  ...A................
K    3510 hCoV-19/England/QEUH-9C7DF6/2020|EPI_ISL_567035|20    ....................  ...A................  ....................  ....................  ....................  ....................
K    3511 hCoV-19/England/QEUH-9C8681/2020|EPI_ISL_567036|20    ....................  ....................  ....................  ....................  ....................  ....................
K    3512 hCoV-19/England/QEUH-9C8AB2/2020|EPI_ISL_567038|20    ....................  ....................  ....................  ....................  ....................  ....................
K    3513 hCoV-19/England/QEUH-9C8900/2020|EPI_ISL_567039|20    ....................  ....................  ....................  ....................  ....................  ....................
K    3514 hCoV-19/Scotland/QEUH-98FCB1/2020|EPI_ISL_568328|2    ....................  ....................  ....................  ....................  ....................  ....................
K    3515 hCoV-19/Scotland/QEUH-998DA3/2020|EPI_ISL_568329|2    ....................  ....................  ....................  ....................  ....................  ....................
K    3516 hCoV-19/Scotland/QEUH-98FF03/2020|EPI_ISL_568330|2    ....................  ....................  ....................  ....................  ....................  ....................
K    3517 hCoV-19/Scotland/QEUH-998A7F/2020|EPI_ISL_568331|2    ....................  ....................  ....................  ....................  ....................  ....................
K    3518 hCoV-19/Scotland/QEUH-99042E/2020|EPI_ISL_568332|2    ....................  ....................  ....................  ....................  ....................  ....................
K    3519 hCoV-19/Scotland/QEUH-990215/2020|EPI_ISL_568333|2    ....................  ....................  ....................  ....................  ....................  ....................
K    3520 hCoV-19/Scotland/QEUH-98F738/2020|EPI_ISL_568334|2    ....................  ....................  ....................  ....................  ....................  ....................
K    3521 hCoV-19/Scotland/QEUH-998DD0/2020|EPI_ISL_568335|2    ....................  ....................  ....................  ....................  ....................  ....................
K    3522 hCoV-19/Scotland/QEUH-98FF12/2020|EPI_ISL_568336|2    ....................  ....................  ....................  ....................  ....................  ....................
K    3523 hCoV-19/Scotland/QEUH-9983A1/2020|EPI_ISL_568337|2    ....................  ....................  ....................  ....................  ....................  ....................
K    3524 hCoV-19/Scotland/QEUH-98F6C2/2020|EPI_ISL_568338|2    ....................  ....................  ....................  ....................  ....................  ....................
K    3525 hCoV-19/Scotland/QEUH-98FA8A/2020|EPI_ISL_568339|2    ....................  ....................  ....................  ....................  ....................  ....................
K    3526 hCoV-19/Scotland/QEUH-998C97/2020|EPI_ISL_568340|2    ....................  ....................  ....................  ....................  ....................  ....................
K    3527 hCoV-19/Scotland/QEUH-98F756/2020|EPI_ISL_568341|2    ....................  ....................  ....................  ....................  ....................  ....................
K    3528 hCoV-19/Scotland/QEUH-98FD18/2020|EPI_ISL_568342|2    ....................  ....................  ....................  ....................  ....................  ....................
K    3529 hCoV-19/Scotland/QEUH-998D1C/2020|EPI_ISL_568343|2    ....................  ....................  ....................  ....................  ....................  ....................
K    3530 hCoV-19/Scotland/QEUH-98F9D8/2020|EPI_ISL_568345|2    ....................  ....................  ....................  ....................  ....................  ....................
K    3531 hCoV-19/Scotland/QEUH-998DFE/2020|EPI_ISL_568346|2    ....................  ....................  ....................  ....................  ....................  ....................
K    3532 hCoV-19/Scotland/QEUH-998D58/2020|EPI_ISL_568347|2    ....................  ....................  ....................  ....................  ....................  ....................
K    3533 hCoV-19/Scotland/QEUH-998B03/2020|EPI_ISL_568348|2    ....................  ....................  ....................  ....................  ....................  ....................
K    3534 hCoV-19/Scotland/QEUH-98F862/2020|EPI_ISL_568349|2    ....................  ....................  ....................  ....................  ....................  ....................
K    3535 hCoV-19/Scotland/QEUH-98F853/2020|EPI_ISL_568359|2    ....................  ....................  ....................  ....................  ....................  ....................
K    3536 hCoV-19/Scotland/QEUH-998C00/2020|EPI_ISL_568360|2    ....................  ....................  ....................  ....................  ....................  ....................
K    3537 hCoV-19/Scotland/QEUH-998ABB/2020|EPI_ISL_568361|2    ....................  ....................  ....................  ....................  ....................  ....................
K    3538 hCoV-19/Scotland/QEUH-9900EE/2020|EPI_ISL_568362|2    ....................  ....................  ....................  ....................  ....................  ....................
K    3539 hCoV-19/Scotland/QEUH-990637/2020|EPI_ISL_568363|2    ....................  ....................  ....................  ....................  ....................  ....................
K    3540 hCoV-19/England/MILK-99E262/2020|EPI_ISL_568364|20    ....................  ....................  ....................  ....................  ....................  ....................
K    3541 hCoV-19/England/MILK-99DF11/2020|EPI_ISL_568365|20    ....................  ....................  ....................  ....................  ....................  ....................
K    3542 hCoV-19/Scotland/QEUH-9990C7/2020|EPI_ISL_568366|2    ....................  .A..................  ....................  ....................  ....................  ....................
K    3543 hCoV-19/Scotland/QEUH-999267/2020|EPI_ISL_568367|2    ....................  ....................  ....................  ....................  ....................  ....................
K    3544 hCoV-19/Scotland/QEUH-9A46D6/2020|EPI_ISL_568368|2    ....................  ....................  ....................  ....................  ....................  ....................
K    3545 hCoV-19/Scotland/QEUH-9A409C/2020|EPI_ISL_568369|2    ....................  ....................  ....................  ....................  ....................  ....................
K    3546 hCoV-19/USA/NM-NMDOH-2020328645/2020|EPI_ISL_56965    ....................  ....................  ....................  ....................  ....................  ....................
K    3547 hCoV-19/USA/NM-NMDOH-2020328767/2020|EPI_ISL_56966    ....................  .A..................  ....................  ....................  ....................  ....................
K    3548 hCoV-19/USA/NM-NMDOH-2020329070/2020|EPI_ISL_56966    ....................  ....................  ....................  ....................  ....................  ....................
K    3549 hCoV-19/USA/NM-NMDOH-2020329281/2020|EPI_ISL_56966    ....................  ....................  ....................  ....................  ....................  ....................
K    3550 hCoV-19/USA/NM-NMDOH-2020329356/2020|EPI_ISL_56966    ....................  .A..................  ....................  ....................  ....................  ....................
K    3551 hCoV-19/USA/WA-UW-22212/2020|EPI_ISL_570640|2020-0    ....................  ....................  ....................  ....................  ....................  ....................
K    3552 hCoV-19/USA/WA-UW-22217/2020|EPI_ISL_570641|2020-0    ....................  ....................  ....................  ....................  ....................  ....................
K    3553 hCoV-19/USA/WA-UW-22228/2020|EPI_ISL_570642|2020-0    ....................  ....................  ....................  ....................  ....................  ....................
K    3554 hCoV-19/USA/WA-UW-22240/2020|EPI_ISL_570643|2020-0    ....................  .A..................  ....................  ....................  ....................  ....................
K    3555 hCoV-19/USA/WA-UW-22247/2020|EPI_ISL_570644|2020-0    ....................  ....................  ....................  ....................  ....................  ....................
K    3556 hCoV-19/USA/WA-UW-22253/2020|EPI_ISL_570645|2020-0    ....................  ....................  ....................  ....................  ....................  ....................
K    3557 hCoV-19/USA/WA-UW-22258/2020|EPI_ISL_570646|2020-0    ....................  ....................  ....................  ....................  ....................  ....................
K    3558 hCoV-19/USA/WA-UW-22273/2020|EPI_ISL_570647|2020-0    ....................  ....................  ....................  ....................  ....................  ....................
K    3559 hCoV-19/USA/WA-UW-22275/2020|EPI_ISL_570648|2020-0    ....................  ....................  ....................  ....................  ....................  ....................
K    3560 hCoV-19/USA/WA-UW-22281/2020|EPI_ISL_570649|2020-0    ....................  ....................  ....................  ....................  ....................  ....................
K    3561 hCoV-19/USA/WA-UW-22286/2020|EPI_ISL_570650|2020-0    ....................  ....................  ....................  ....................  ....................  ....................
K    3562 hCoV-19/USA/WA-UW-22287/2020|EPI_ISL_570651|2020-0    ....................  ....................  ....................  ....................  ....................  ....................
K    3563 hCoV-19/USA/WA-UW-22293/2020|EPI_ISL_570652|2020-0    ....................  ....................  ....................  ....................  ....................  ....................
K    3564 hCoV-19/USA/WA-UW-22294/2020|EPI_ISL_570653|2020-0    ....................  ....................  ....................  ....................  ....................  ....................
K    3565 hCoV-19/USA/WA-UW-22302/2020|EPI_ISL_570654|2020-0    ....................  ....................  ....................  ....................  ....................  ....................
K    3566 hCoV-19/USA/WA-UW-22306/2020|EPI_ISL_570655|2020-0    ....................  ....................  ....................  ....................  ....................  ....................
K    3567 hCoV-19/USA/WA-UW-22312/2020|EPI_ISL_570656|2020-0    ....................  ....................  ....................  ....................  ....................  ....................
K    3568 hCoV-19/USA/WA-UW-22313/2020|EPI_ISL_570657|2020-0    ....................  .A..................  ....................  ....................  ....................  ....................
K    3569 hCoV-19/USA/WA-UW-22316/2020|EPI_ISL_570658|2020-0    ....................  .A..................  ....................  ....................  ....................  ....................
K    3570 hCoV-19/USA/WA-UW-22318/2020|EPI_ISL_570659|2020-0    ....................  ....................  ....................  ....................  ....................  ....................
K    3571 hCoV-19/USA/WA-UW-22363/2020|EPI_ISL_570668|2020-0    ....................  ....................  ....................  ....................  ....................  ....................
K    3572 hCoV-19/USA/WA-UW-22365/2020|EPI_ISL_570669|2020-0    ....................  .A..................  ....................  ....................  ....................  ....................
K    3573 hCoV-19/USA/WA-UW-22370/2020|EPI_ISL_570670|2020-0    ....................  ....................  ....................  ....................  ....................  ....................
K    3574 hCoV-19/USA/WA-UW-22376/2020|EPI_ISL_570671|2020-0    ....................  ....................  ....................  ....................  ....................  ....................
K    3575 hCoV-19/USA/WA-UW-22382/2020|EPI_ISL_570672|2020-0    ....................  ....................  ....................  ....................  ....................  ....................
K    3576 hCoV-19/USA/WA-UW-22389/2020|EPI_ISL_570673|2020-0    ....................  ....................  ....................  ....................  ....................  ....................
K    3577 hCoV-19/USA/WA-UW-22398/2020|EPI_ISL_570674|2020-0    ....................  .A..................  ....................  ....................  ....................  ....................
K    3578 hCoV-19/USA/WA-UW-22399/2020|EPI_ISL_570675|2020-0    ....................  ....................  ....................  ....................  ....................  ....................
K    3579 hCoV-19/USA/WA-UW-22405/2020|EPI_ISL_570676|2020-0    ....................  ....................  ....................  ....................  ....................  ....................
K    3580 hCoV-19/USA/WA-UW-22407/2020|EPI_ISL_570677|2020-0    ....................  .A..................  ....................  ....................  ....................  ....................
K    3581 hCoV-19/USA/WA-UW-22409/2020|EPI_ISL_570678|2020-0    ....................  .A..................  ....................  ....................  ....................  ....................
K    3582 hCoV-19/USA/WA-UW-22412/2020|EPI_ISL_570679|2020-0    ....................  ....................  ....................  ....................  ....................  ....................
K    3583 hCoV-19/USA/WA-UW-22418/2020|EPI_ISL_570680|2020-0    ....................  ....................  ....................  ....................  ....................  ....................
K    3584 hCoV-19/USA/WA-UW-22419/2020|EPI_ISL_570681|2020-0    ....................  ....................  ....................  ....................  ....................  ....................
K    3585 hCoV-19/Australia/VIC12356/2020|EPI_ISL_563554|202    ....................  ....................  ....................  ....................  ....................  ....................
K    3586 hCoV-19/Australia/VIC12361/2020|EPI_ISL_563555|202    ....................  ....................  ....................  ....................  ....................  ....................
K    3587 hCoV-19/Australia/VIC12380/2020|EPI_ISL_563556|202    ....................  ....................  ....................  ....................  ....................  ....................
K    3588 hCoV-19/Australia/VIC12385/2020|EPI_ISL_563557|202    ....................  ....................  ....................  ....................  ....................  ....................
```

| | | | | | | | |
|---|---|---|---|---|---|---|---|
| K | 3589 hCoV-19/Australia/VIC12389/2020\|EPI_ISL_563558\|202 | .................... | .................... | .................... | .................... | .................... | .................... |
| K | 3590 hCoV-19/Australia/VIC12400/2020\|EPI_ISL_563559\|202 | .................... | .................... | .................... | .................... | .................... | .................... |
| K | 3591 hCoV-19/Australia/VIC12407/2020\|EPI_ISL_563560\|202 | .................... | .................... | .................... | .................... | .................... | .................... |
| K | 3592 hCoV-19/Australia/VIC12410/2020\|EPI_ISL_563561\|202 | .................... | .................... | .................... | .................... | .................... | .................... |
| K | 3593 hCoV-19/Australia/VIC12418/2020\|EPI_ISL_563562\|202 | .................... | .................... | .................... | .................... | .................... | .................... |
| K | 3594 hCoV-19/Australia/VIC12438/2020\|EPI_ISL_563563\|202 | .................... | .................... | .................... | .................... | .................... | .................... |
| K | 3595 hCoV-19/Australia/VIC12449/2020\|EPI_ISL_563564\|202 | .................... | .................... | .................... | .................... | .................... | .................... |
| K | 3596 hCoV-19/Australia/VIC12453/2020\|EPI_ISL_563565\|202 | .................... | .................... | .................... | .................... | .................... | .................... |
| K | 3597 hCoV-19/Australia/VIC12476/2020\|EPI_ISL_563566\|202 | .................... | .................... | .................... | .................... | .................... | .................... |
| K | 3598 hCoV-19/Australia/VIC12484/2020\|EPI_ISL_563567\|202 | .................... | .................... | .................... | .................... | .................... | .................... |
| K | 3599 hCoV-19/Australia/VIC12524/2020\|EPI_ISL_563568\|202 | .................... | .................... | .................... | .................... | .................... | .................... |
| K | 3600 hCoV-19/Australia/VIC12526/2020\|EPI_ISL_563569\|202 | .................... | .................... | .................... | .................... | .................... | .................... |
| K | 3601 hCoV-19/Australia/VIC12535/2020\|EPI_ISL_563570\|202 | .................... | .................... | .................... | .................... | .................... | .................... |
| K | 3602 hCoV-19/Australia/VIC12551/2020\|EPI_ISL_563571\|202 | .................... | .................... | .................... | .................... | .................... | .................... |
| K | 3603 hCoV-19/Australia/VIC12555/2020\|EPI_ISL_563572\|202 | .................... | .................... | .................... | .................... | .................... | .................... |
| K | 3604 hCoV-19/Australia/VIC12585/2020\|EPI_ISL_563573\|202 | .................... | .................... | .................... | .................... | .................... | .................... |
| K | 3605 hCoV-19/Australia/VIC12591/2020\|EPI_ISL_563574\|202 | .................... | .................... | .................... | .................... | .................... | .................... |
| K | 3606 hCoV-19/Australia/VIC12597/2020\|EPI_ISL_563575\|202 | .................... | .................... | .................... | .................... | .................... | .................... |
| K | 3607 hCoV-19/Australia/VIC12743/2020\|EPI_ISL_563587\|202 | .................... | .................... | .................... | .................... | .................... | .................... |
| K | 3608 hCoV-19/Australia/VIC12750/2020\|EPI_ISL_563588\|202 | .................... | .................... | .................... | .................... | .................... | .................... |
| K | 3609 hCoV-19/Australia/VIC12756/2020\|EPI_ISL_563589\|202 | .................... | .................... | .................... | .................... | .................... | .................... |
| K | 3611 hCoV-19/Australia/VIC12796/2020\|EPI_ISL_563595\|202 | .................... | .................... | .................... | .................... | .................... | .................... |
| K | 3612 hCoV-19/Australia/VIC8175/2020\|EPI_ISL_564885\|2020 | .................... | .................... | .................... | .................... | .................... | .................... |
| K | 3613 hCoV-19/Australia/VIC8181/2020\|EPI_ISL_564886\|2020 | .................... | .................... | .................... | .................... | .................... | .................... |
| K | 3614 hCoV-19/Australia/VIC8182/2020\|EPI_ISL_564887\|2020 | .................... | .................... | .................... | .................... | .................... | .................... |
| K | 3615 hCoV-19/Australia/VIC8183/2020\|EPI_ISL_564888\|2020 | .................... | .................... | .................... | .................... | .................... | .................... |
| K | 3616 hCoV-19/Australia/VIC8184/2020\|EPI_ISL_564889\|2020 | .................... | .................... | .................... | .................... | .................... | .................... |
| K | 3617 hCoV-19/Australia/VIC8187/2020\|EPI_ISL_564890\|2020 | .................... | .................... | .................... | .................... | .................... | .................... |
| K | 3618 hCoV-19/Australia/VIC8189/2020\|EPI_ISL_564891\|2020 | .................... | .................... | .................... | .................... | .................... | .................... |
| K | 3619 hCoV-19/Australia/VIC8191/2020\|EPI_ISL_564892\|2020 | .................... | .................... | .................... | .................... | .................... | .................... |
| K | 3620 hCoV-19/Australia/VIC8195/2020\|EPI_ISL_564893\|2020 | .................... | .................... | .................... | .................... | .................... | .................... |
| K | 3621 hCoV-19/Australia/VIC8196/2020\|EPI_ISL_564894\|2020 | .................... | .................... | .................... | .................... | .................... | .................... |
| K | 3622 hCoV-19/Australia/VIC8197/2020\|EPI_ISL_564895\|2020 | .................... | .................... | .................... | .................... | .................... | .................... |
| K | 3623 hCoV-19/Australia/VIC8199/2020\|EPI_ISL_564896\|2020 | .................... | .................... | .................... | .................... | .................... | .................... |
| K | 3624 hCoV-19/Australia/VIC8202/2020\|EPI_ISL_564897\|2020 | .................... | .................... | .................... | .................... | .................... | .................... |
| K | 3625 hCoV-19/Australia/VIC8203/2020\|EPI_ISL_564898\|2020 | .................... | .................... | .................... | .................... | .................... | .................... |
| K | 3626 hCoV-19/Australia/VIC8207/2020\|EPI_ISL_564899\|2020 | .................... | .................... | .................... | .................... | .................... | .................... |
| K | 3627 hCoV-19/England/QEUH-9C84E1/2020\|EPI_ISL_567020\|20 | .................... | .................... | .................... | .................... | .................... | .................... |
| K | 3628 hCoV-19/England/QEUH-9C90FB/2020\|EPI_ISL_567021\|20 | .................... | .................... | .................... | .................... | .................... | .................... |
| K | 3629 hCoV-19/England/QEUH-9C80B0/2020\|EPI_ISL_567022\|20 | .................... | .................... | .................... | .................... | .................... | .................... |
| K | 3630 hCoV-19/England/QEUH-9C838A/2020\|EPI_ISL_567023\|20 | .................... | .................... | .................... | .................... | .................... | .................... |
| K | 3631 hCoV-19/England/QEUH-9C844B/2020\|EPI_ISL_567025\|20 | .................... | .................... | .................... | .................... | .................... | .................... |
| K | 3632 hCoV-19/England/QEUH-9C8D04/2020\|EPI_ISL_567026\|20 | .................... | .................... | .................... | .................... | .................... | .................... |
| K | 3633 hCoV-19/England/QEUH-9C852A/2020\|EPI_ISL_567028\|20 | .................... | .................... | .................... | .................... | .....T.............. | .................... |
| K | 3634 hCoV-19/England/QEUH-9C8AFE/2020\|EPI_ISL_567029\|20 | .................... | .A.................. | .................... | .................... | .................... | ...A................ |
| K | 3635 hCoV-19/England/QEUH-9C7C08/2020\|EPI_ISL_567040\|20 | .................... | .................... | .................... | .................... | .................... | .................... |
| K | 3636 hCoV-19/England/QEUH-9C7C26/2020\|EPI_ISL_567041\|20 | .................... | .................... | .................... | .................... | .................... | .................... |
| K | 3637 hCoV-19/England/QEUH-9C896A/2020\|EPI_ISL_567042\|20 | .................... | .................... | .................... | .................... | .................... | .................... |
| K | 3638 hCoV-19/England/QEUH-9C7FB4/2020\|EPI_ISL_567044\|20 | .................... | .................... | .................... | .................... | .................... | .................... |
| K | 3639 hCoV-19/England/QEUH-9C8B46/2020\|EPI_ISL_567045\|20 | .................... | .................... | .................... | .................... | .................... | .................... |
| K | 3640 hCoV-19/England/QEUH-9C8214/2020\|EPI_ISL_567046\|20 | .................... | .................... | .................... | .................... | .................... | .................... |
| K | 3641 hCoV-19/England/QEUH-9C8C9E/2020\|EPI_ISL_567047\|20 | .................... | .................... | .................... | .................... | .................... | .................... |
| K | 3642 hCoV-19/England/QEUH-9C8557/2020\|EPI_ISL_567049\|20 | .................... | .A.................. | .................... | .................... | .................... | .................... |
| K | 3643 hCoV-19/England/QEUH-9C7EA8/2020\|EPI_ISL_567050\|20 | .................... | .................... | .................... | .................... | .................... | .................... |
| K | 3644 hCoV-19/England/QEUH-9C91F8/2020\|EPI_ISL_567051\|20 | .................... | .................... | .................... | .................... | .................... | .................... |
| K | 3645 hCoV-19/England/QEUH-9C7B38/2020\|EPI_ISL_567052\|20 | .................... | .................... | .................... | .................... | .................... | .................... |
| K | 3646 hCoV-19/England/QEUH-9C919E/2020\|EPI_ISL_567053\|20 | .................... | .................... | .................... | .................... | .................... | .................... |
| K | 3647 hCoV-19/England/QEUH-9C8AA3/2020\|EPI_ISL_567054\|20 | .................... | .................... | .................... | .................... | .................... | .................... |
| K | 3648 hCoV-19/England/QEUH-9C9064/2020\|EPI_ISL_567055\|20 | .................... | .................... | .................... | .................... | .................... | .................... |
| K | 3649 hCoV-19/England/QEUH-9C7E11/2020\|EPI_ISL_567056\|20 | .................... | .................... | .................... | .................... | .................... | .................... |
| K | 3650 hCoV-19/England/QEUH-9C8DD7/2020\|EPI_ISL_567057\|20 | .................... | .................... | .................... | .................... | .................... | .................... |
| K | 3651 hCoV-19/England/QEUH-9C7FE1/2020\|EPI_ISL_567058\|20 | .................... | .................... | .................... | .................... | .................... | .................... |
| K | 3652 hCoV-19/England/QEUH-9C82AB/2020\|EPI_ISL_567060\|20 | .................... | .................... | .................... | .................... | .................... | .................... |
| K | 3653 hCoV-19/England/QEUH-9C7ED5/2020\|EPI_ISL_567061\|20 | .................... | .................... | .................... | .................... | .................... | .................... |
| K | 3654 hCoV-19/England/QEUH-9C8241/2020\|EPI_ISL_567062\|20 | .................... | .................... | .................... | .................... | .................... | .................... |
| K | 3655 hCoV-19/England/QEUH-9C8D6E/2020\|EPI_ISL_567063\|20 | .................... | .................... | .................... | .................... | .................... | .................... |
| K | 3656 hCoV-19/England/QEUH-9C8997/2020\|EPI_ISL_567064\|20 | .................... | .................... | .................... | .................... | .................... | .................... |
| K | 3657 hCoV-19/England/QEUH-9C887C/2020\|EPI_ISL_567065\|20 | .................... | .................... | .................... | .................... | .................... | .................... |
| K | 3658 hCoV-19/England/QEUH-9C7AE0/2020\|EPI_ISL_567066\|20 | .................... | .................... | .................... | .................... | .................... | .................... |
| K | 3659 hCoV-19/England/QEUH-9C81EA/2020\|EPI_ISL_567067\|20 | .................... | .................... | .................... | .................... | .................... | .................... |
| K | 3660 hCoV-19/England/QEUH-9C827E/2020\|EPI_ISL_567068\|20 | .................... | .A.................. | .................... | .................... | .................... | .................... |
| K | 3661 hCoV-19/England/QEUH-9C7E5D/2020\|EPI_ISL_567069\|20 | .................... | .................... | .................... | .................... | .................... | .................... |
| K | 3662 hCoV-19/England/QEUH-9C8584/2020\|EPI_ISL_567070\|20 | .................... | .................... | .................... | .................... | .................... | .................... |
| K | 3663 hCoV-19/England/QEUH-9C8320/2020\|EPI_ISL_567072\|20 | .................... | .................... | .................... | .................... | .................... | .................... |
| K | 3664 hCoV-19/England/QEUH-9C841E/2020\|EPI_ISL_567073\|20 | .................... | .................... | .................... | .................... | .................... | .................... |
| K | 3665 hCoV-19/England/QEUH-9C7DE7/2020\|EPI_ISL_567074\|20 | .................... | .................... | .................... | .................... | .....T.............. | .................... |
| K | 3666 hCoV-19/England/QEUH-9C7D8D/2020\|EPI_ISL_567075\|20 | .................... | .................... | .................... | .................... | .................... | .................... |
| K | 3667 hCoV-19/England/QEUH-9C86DC/2020\|EPI_ISL_567076\|20 | .................... | .................... | .................... | .................... | .................... | .................... |
| K | 3668 hCoV-19/England/QEUH-9C82E7/2020\|EPI_ISL_567078\|20 | .................... | .................... | .................... | .................... | .................... | .................... |
| K | 3669 hCoV-19/England/QEUH-9C84D2/2020\|EPI_ISL_567079\|20 | .................... | .................... | .................... | .................... | .................... | .................... |

| | | | | | | | | |
|---|---|---|---|---|---|---|---|---|
| K | 3670 | hCoV-19/Scotland/QEUH-998E37/2020\|EPI_ISL_568302\|2 | ···················· | ···················· | ····················· | ···················· | ···················· | ···················· |
| K | 3671 | hCoV-19/Scotland/QEUH-98FE9D/2020\|EPI_ISL_568304\|2 | ···················· | ···················· | ····················· | ···················· | ···················· | ···················· |
| K | 3672 | hCoV-19/Scotland/QEUH-998ACA/2020\|EPI_ISL_568305\|2 | ···················· | ···················· | ····················· | ···················· | ···················· | ···················· |
| K | 3673 | hCoV-19/Scotland/QEUH-99043D/2020\|EPI_ISL_568306\|2 | ···················· | ···················· | ····················· | ···················· | ···················· | ···················· |
| K | 3674 | hCoV-19/Scotland/QEUH-990224/2020\|EPI_ISL_568307\|2 | ···················· | ···················· | ····················· | ···················· | ···················· | ···················· |
| K | 3675 | hCoV-19/Scotland/QEUH-98FA7B/2020\|EPI_ISL_568308\|2 | ···················· | ···················· | ····················· | ···················· | ···················· | ···················· |
| K | 3676 | hCoV-19/Scotland/QEUH-998B8B/2020\|EPI_ISL_568309\|2 | ···················· | ···················· | ····················· | ···················· | ···················· | ···················· |
| K | 3677 | hCoV-19/Scotland/QEUH-998DB2/2020\|EPI_ISL_568310\|2 | ···················· | ···················· | ····················· | ···················· | ···················· | ···················· |
| K | 3678 | hCoV-19/Scotland/QEUH-98FAD5/2020\|EPI_ISL_568311\|2 | ···················· | ···················· | ····················· | ···················· | ···················· | ···················· |
| K | 3679 | hCoV-19/Scotland/QEUH-998D3A/2020\|EPI_ISL_568312\|2 | ···················· | ···················· | ····················· | ···················· | ···················· | ···················· |
| K | 3680 | hCoV-19/Scotland/QEUH-998B7C/2020\|EPI_ISL_568313\|2 | ···················· | ···················· | ····················· | ···················· | ···················· | ···················· |
| K | 3681 | hCoV-19/Scotland/QEUH-98FD27/2020\|EPI_ISL_568314\|2 | ···················· | ···················· | ····················· | ···················· | ···················· | ···················· |
| K | 3682 | hCoV-19/Scotland/QEUH-98F60E/2020\|EPI_ISL_568316\|2 | ···················· | ···················· | ····················· | ···················· | ···················· | ···················· |
| K | 3683 | hCoV-19/Scotland/QEUH-998DEF/2020\|EPI_ISL_568318\|2 | ···················· | ···················· | ····················· | ···················· | ···················· | ···················· |
| K | 3684 | hCoV-19/Scotland/QEUH-998D0D/2020\|EPI_ISL_568319\|2 | ···················· | ···················· | ····················· | ···················· | ···················· | ···················· |
| K | 3685 | hCoV-19/Scotland/QEUH-998CF1/2020\|EPI_ISL_568320\|2 | ···················· | ···················· | ····················· | ···················· | ···················· | ···················· |
| K | 3686 | hCoV-19/Scotland/QEUH-98F45F/2020\|EPI_ISL_568321\|2 | ···················· | ···················· | ····················· | ···················· | ···················· | ···················· |
| K | 3687 | hCoV-19/Scotland/QEUH-98F598/2020\|EPI_ISL_568322\|2 | ···················· | ···················· | ····················· | ···················· | ···················· | ···················· |
| K | 3688 | hCoV-19/Scotland/QEUH-99034F/2020\|EPI_ISL_568323\|2 | ···················· | ···················· | ····················· | ···················· | ···················· | ···················· |
| K | 3689 | hCoV-19/Scotland/QEUH-98FFC7/2020\|EPI_ISL_568324\|2 | ···················· | ···················· | ····················· | ···················· | ···················· | ···················· |
| K | 3690 | hCoV-19/Scotland/QEUH-98FB2D/2020\|EPI_ISL_568326\|2 | ···················· | ···················· | ····················· | ···················· | ······T············· | ···················· |
| K | 3691 | hCoV-19/Scotland/QEUH-9901CD/2020\|EPI_ISL_568327\|2 | ···················· | ···················· | ····················· | ···················· | ···················· | ···················· |
| K | 3692 | hCoV-19/Scotland/QEUH-998D85/2020\|EPI_ISL_568350\|2 | ···················· | ···················· | ····················· | ···················· | ···················· | ···················· |
| K | 3693 | hCoV-19/Scotland/QEUH-98FB5A/2020\|EPI_ISL_568351\|2 | ···················· | ···················· | ····················· | ···················· | ···················· | ···················· |
| K | 3694 | hCoV-19/Scotland/QEUH-998A8E/2020\|EPI_ISL_568352\|2 | ···················· | ···················· | ····················· | ···················· | ···················· | ···················· |
| K | 3695 | hCoV-19/Scotland/QEUH-998347/2020\|EPI_ISL_568353\|2 | ···················· | ···················· | ····················· | ···················· | ···················· | ···················· |
| K | 3696 | hCoV-19/Scotland/QEUH-9983B0/2020\|EPI_ISL_568354\|2 | ···················· | ···················· | ····················· | ···················· | ···················· | ···················· |
| K | 3697 | hCoV-19/Scotland/QEUH-998374/2020\|EPI_ISL_568355\|2 | ···················· | ·A·················· | ····················· | ···················· | ···················· | ···A················ |
| K | 3698 | hCoV-19/Scotland/QEUH-98F96F/2020\|EPI_ISL_568357\|2 | ···················· | ···················· | ····················· | ···················· | ···················· | ···················· |
| K | 3699 | hCoV-19/Scotland/QEUH-98F765/2020\|EPI_ISL_568358\|2 | ···················· | ···················· | ····················· | ···················· | ···················· | ···················· |
| K | 3700 | hCoV-19/Scotland/QEUH-9A476A/2020\|EPI_ISL_568370\|2 | ···················· | ···················· | ····················· | ···················· | ···················· | ···················· |
| K | 3701 | hCoV-19/England/QEUH-9A473D/2020\|EPI_ISL_568371\|20 | ···················· | ···················· | ····················· | ···················· | ···················· | ···················· |
| K | 3702 | hCoV-19/Scotland/QEUH-9A41D5/2020\|EPI_ISL_568372\|2 | ···················· | ···················· | ····················· | ···················· | ···················· | ···················· |
| K | 3703 | hCoV-19/Scotland/QEUH-9A41F3/2020\|EPI_ISL_568373\|2 | ···················· | ···················· | ····················· | ···················· | ···················· | ···················· |
| K | 3704 | hCoV-19/Scotland/QEUH-9A46B8/2020\|EPI_ISL_568374\|2 | ···················· | ···················· | ····················· | ···················· | ···················· | ···················· |
| K | 3705 | hCoV-19/Scotland/QEUH-9A4779/2020\|EPI_ISL_568375\|2 | ···················· | ···················· | ····················· | ···················· | ···················· | ···················· |
| K | 3706 | hCoV-19/Scotland/QEUH-9A4050/2020\|EPI_ISL_568376\|2 | ···················· | ···················· | ····················· | ···················· | ···················· | ···················· |
| K | 3707 | hCoV-19/Scotland/QEUH-9A45F7/2020\|EPI_ISL_568377\|2 | ···················· | ···················· | ····················· | ···················· | ···················· | ···················· |
| K | 3708 | hCoV-19/Scotland/QEUH-9A4041/2020\|EPI_ISL_568378\|2 | ···················· | ···················· | ····················· | ···················· | ···················· | ···················· |
| K | 3709 | hCoV-19/Scotland/QEUH-9A4366/2020\|EPI_ISL_568379\|2 | ···················· | ···················· | ····················· | ···················· | ···················· | ···················· |
| K | 3710 | hCoV-19/Scotland/QEUH-9A3FCD/2020\|EPI_ISL_568380\|2 | ···················· | ···················· | ····················· | ···················· | ···················· | ···················· |
| K | 3711 | hCoV-19/Scotland/QEUH-9A474C/2020\|EPI_ISL_568381\|2 | ···················· | ···················· | ····················· | ···················· | ···················· | ···················· |
| K | 3712 | hCoV-19/Scotland/QEUH-9A44CD/2020\|EPI_ISL_568382\|2 | ···················· | ···················· | ····················· | ···················· | ···················· | ···················· |
| K | 3713 | hCoV-19/Scotland/QEUH-9A472E/2020\|EPI_ISL_568383\|2 | ···················· | ···················· | ····················· | ···················· | ···················· | ···················· |
| K | 3714 | hCoV-19/Scotland/QEUH-96A26F/2020\|EPI_ISL_568384\|2 | ···················· | ···················· | ····················· | ···················· | ···················· | ···················· |
| K | 3715 | hCoV-19/Scotland/CAMC-9914C3/2020\|EPI_ISL_568385\|2 | ···················· | ···················· | ····················· | ···················· | ···················· | ···················· |
| K | 3716 | hCoV-19/Scotland/MILK-993C8D/2020\|EPI_ISL_568386\|2 | ···················· | ···················· | ····················· | ···················· | ···················· | ···················· |
| K | 3717 | hCoV-19/Scotland/MILK-994D3E/2020\|EPI_ISL_568387\|2 | ···················· | ···················· | ····················· | ···················· | ···················· | ···················· |
| K | 3718 | hCoV-19/Scotland/QEUH-997EFC/2020\|EPI_ISL_568388\|2 | ···················· | ···················· | ····················· | ···················· | ···················· | ···················· |
| K | 3719 | hCoV-19/Scotland/QEUH-998110/2020\|EPI_ISL_568389\|2 | ···················· | ···················· | ····················· | ···················· | ···················· | ···················· |
| K | 3720 | hCoV-19/England/QEUH-99813E/2020\|EPI_ISL_568390\|20 | ···················· | ···················· | ····················· | ···················· | ···················· | ···················· |
| K | 3721 | hCoV-19/Scotland/QEUH-997F80/2020\|EPI_ISL_568391\|2 | ···················· | ···················· | ····················· | ···················· | ···················· | ···················· |
| K | 3722 | hCoV-19/Scotland/QEUH-997D4A/2020\|EPI_ISL_568393\|2 | ···················· | ···················· | ····················· | ···················· | ···················· | ···················· |
| K | 3723 | hCoV-19/England/QEUH-998198/2020\|EPI_ISL_568394\|20 | ···················· | ···················· | ····················· | ···················· | ···················· | ···················· |
| K | 3724 | hCoV-19/France/PAC-IHU-0578/2020\|EPI_ISL_569600\|20 | ···················· | ···················· | ····················· | ···················· | ···················· | ···················· |
| K | 3725 | hCoV-19/France/PAC-IHU-0580/2020\|EPI_ISL_569601\|20 | ···················· | ···················· | ····················· | ···················· | ···················· | ···················· |
| K | 3726 | hCoV-19/France/PAC-IHU-0583/2020\|EPI_ISL_569602\|20 | ···················· | ···················· | ····················· | ···················· | ···················· | ···················· |
| K | 3727 | hCoV-19/France/PAC-IHU-0584/2020\|EPI_ISL_569603\|20 | ···················· | ···················· | ····················· | ···················· | ···················· | ···················· |
| K | 3728 | hCoV-19/France/PAC-IHU-0586/2020\|EPI_ISL_569604\|20 | ···················· | ···················· | ····················· | ···················· | ···················· | ···················· |
| K | 3729 | hCoV-19/France/PAC-IHU-0587/2020\|EPI_ISL_569605\|20 | ···················· | ···················· | ····················· | ···················· | ···················· | ···················· |
| K | 3730 | hCoV-19/France/PAC-IHU-0589/2020\|EPI_ISL_569606\|20 | ···················· | ···················· | ····················· | ···················· | ···················· | ···················· |
| K | 3731 | hCoV-19/France/PAC-IHU-0590/2020\|EPI_ISL_569607\|20 | ···················· | ···················· | ····················· | ···················· | ···················· | ···················· |
| K | 3732 | hCoV-19/France/PAC-IHU-0592/2020\|EPI_ISL_569608\|20 | ···················· | ···················· | ····················· | ···················· | ···················· | ···················· |
| K | 3733 | hCoV-19/USA/SD-SDPHL-0001/2020\|EPI_ISL_569609\|2020 | ···················· | ···················· | ····················· | ···················· | ···················· | ···················· |
| K | 3734 | hCoV-19/USA/SD-SDPHL-0002/2020\|EPI_ISL_569610\|2020 | ···················· | ···················· | ····················· | ···················· | ···················· | ···················· |
| K | 3735 | hCoV-19/USA/SD-SDPHL-0003/2020\|EPI_ISL_569611\|2020 | ···················· | ···················· | ····················· | ···················· | ···················· | ···················· |
| K | 3736 | hCoV-19/USA/SD-SDPHL-0004/2020\|EPI_ISL_569612\|2020 | ···················· | ···················· | ····················· | ···················· | ···················· | ···················· |
| K | 3737 | hCoV-19/USA/SD-SDPHL-0005/2020\|EPI_ISL_569613\|2020 | ···················· | ···················· | ····················· | ···················· | ···················· | ···················· |
| K | 3738 | hCoV-19/USA/SD-SDPHL-0006/2020\|EPI_ISL_569614\|2020 | ···················· | ···················· | ····················· | ···················· | ···················· | ···················· |
| K | 3739 | hCoV-19/USA/SD-SDPHL-0008/2020\|EPI_ISL_569616\|2020 | ···················· | ···················· | ····················· | ···················· | ···················· | ···················· |
| K | 3740 | hCoV-19/USA/SD-SDPHL-0009/2020\|EPI_ISL_569617\|2020 | ···················· | ···················· | ····················· | ···················· | ···················· | ···················· |
| K | 3741 | hCoV-19/USA/SD-SDPHL-0010/2020\|EPI_ISL_569618\|2020 | ···················· | ···················· | ····················· | ···················· | ···················· | ···················· |
| K | 3742 | hCoV-19/USA/SD-SDPHL-0011/2020\|EPI_ISL_569619\|2020 | ···················· | ···················· | ····················· | ···················· | ···················· | ···················· |
| K | 3743 | hCoV-19/USA/SD-SDPHL-0012/2020\|EPI_ISL_569620\|2020 | ···················· | ···················· | ····················· | ···················· | ···················· | ···················· |
| K | 3744 | hCoV-19/USA/SD-SDPHL-0013/2020\|EPI_ISL_569621\|2020 | ···················· | ···················· | ····················· | ···················· | ···················· | ···················· |
| K | 3745 | hCoV-19/USA/SD-SDPHL-0014/2020\|EPI_ISL_569622\|2020 | ···················· | ····A··············· | ····················· | ···················· | ···················· | ···················· |
| K | 3746 | hCoV-19/USA/SD-SDPHL-0015/2020\|EPI_ISL_569623\|2020 | ···················· | ·A·················· | ····················· | ···················· | ···················· | ···················· |
| K | 3747 | hCoV-19/USA/SD-SDPHL-0016/2020\|EPI_ISL_569624\|2020 | ···················· | ···················· | ····················· | ···················· | ···················· | ···················· |
| K | 3748 | hCoV-19/USA/NM-NMDOH-2020321214/2020\|EPI_ISL_56963 | ···················· | ···················· | ····················· | ···················· | ···················· | ···················· |
| K | 3749 | hCoV-19/USA/NM-NMDOH-2020321219/2020\|EPI_ISL_56963 | ···················· | ···················· | ····················· | ···················· | ···················· | ···················· |

| | | | | | | | | |
|---|---|---|---|---|---|---|---|---|
| K | 3750 | hCoV-19/USA/NM-NMDOH-2020321223/2020\|EPI_ISL_56963 | ···················· | ···················· | ····················· | ···················· | ···················· | ···················· |
| K | 3751 | hCoV-19/USA/NM-NMDOH-2020322589/2020\|EPI_ISL_56963 | ···················· | ···················· | ····················· | ···················· | ···················· | ···················· |
| K | 3752 | hCoV-19/USA/NM-NMDOH-2020322592/2020\|EPI_ISL_56963 | ···················· | ···················· | ····················· | ···················· | ···················· | ···················· |
| K | 3753 | hCoV-19/USA/NM-NMDOH-2020324092/2020\|EPI_ISL_56963 | ···················· | ···················· | ····················· | ···················· | ···················· | ···················· |
| K | 3754 | hCoV-19/USA/NM-NMDOH-2020324349/2020\|EPI_ISL_56963 | ···················· | ···················· | ····················· | ···················· | ···················· | ···················· |
| K | 3755 | hCoV-19/USA/NM-NMDOH-2020324378/2020\|EPI_ISL_56963 | ···················· | ···················· | ····················· | ···················· | ···················· | ···················· |
| K | 3756 | hCoV-19/USA/NM-NMDOH-2020326789/2020\|EPI_ISL_56964 | ···················· | ···················· | ····················· | ···················· | ···················· | ···················· |
| K | 3757 | hCoV-19/USA/NM-NMDOH-2020326823/2020\|EPI_ISL_56964 | ···················· | ···················· | ····················· | ···················· | ···················· | ···················· |
| K | 3758 | hCoV-19/USA/NM-NMDOH-2020327022/2020\|EPI_ISL_56964 | ···················· | ·A·················· | ····················· | ···················· | ···················· | ···················· |
| K | 3759 | hCoV-19/USA/NM-NMDOH-2020327026/2020\|EPI_ISL_56964 | ···················· | ·A·················· | ····················· | ···················· | ···················· | ···················· |
| K | 3760 | hCoV-19/USA/NM-NMDOH-2020327064/2020\|EPI_ISL_56964 | ···················· | ···················· | ····················· | ···················· | ···················· | ···················· |
| K | 3761 | hCoV-19/USA/NM-NMDOH-2020327107/2020\|EPI_ISL_56964 | ···················· | ···················· | ····················· | ···················· | ···················· | ···················· |
| K | 3762 | hCoV-19/USA/NM-NMDOH-2020327163/2020\|EPI_ISL_56964 | ···················· | ···················· | ····················· | ···················· | ···················· | ···················· |
| K | 3763 | hCoV-19/USA/NM-NMDOH-2020327257/2020\|EPI_ISL_56964 | ···················· | ···················· | ····················· | ···················· | ···················· | ···················· |
| K | 3764 | hCoV-19/USA/NM-NMDOH-2020327510/2020\|EPI_ISL_56964 | ···················· | ···················· | ····················· | ···················· | ···················· | ···················· |
| K | 3765 | hCoV-19/USA/NM-NMDOH-2020327831/2020\|EPI_ISL_56964 | ···················· | ···················· | ····················· | ···················· | ···················· | ···················· |
| K | 3766 | hCoV-19/USA/NM-NMDOH-2020327863/2020\|EPI_ISL_56965 | ···················· | ···················· | ····················· | ···················· | ···················· | ···················· |
| K | 3767 | hCoV-19/USA/NM-NMDOH-2020328229/2020\|EPI_ISL_56965 | ···················· | ···················· | ····················· | ···················· | ···················· | ···················· |
| K | 3768 | hCoV-19/USA/NM-NMDOH-2020328244/2020\|EPI_ISL_56965 | ···················· | ···················· | ····················· | ···················· | ···················· | ···················· |
| K | 3769 | hCoV-19/USA/NM-NMDOH-2020328404/2020\|EPI_ISL_56965 | ···················· | ···················· | ····················· | ···················· | ···················· | ···················· |
| K | 3770 | hCoV-19/USA/NM-NMDOH-2020328407/2020\|EPI_ISL_56965 | ···················· | ···················· | ····················· | ···················· | ···················· | ···················· |
| K | 3771 | hCoV-19/USA/NM-NMDOH-2020328416/2020\|EPI_ISL_56965 | ···················· | ···················· | ····················· | ···················· | ···················· | ···················· |
| K | 3772 | hCoV-19/USA/NM-NMDOH-2020328523/2020\|EPI_ISL_56965 | ···················· | ···················· | ····················· | ···················· | ···················· | ···················· |
| K | 3773 | hCoV-19/USA/NM-NMDOH-2020328525/2020\|EPI_ISL_56965 | ···················· | ···················· | ····················· | ···················· | ···················· | ···················· |
| K | 3774 | hCoV-19/USA/NM-NMDOH-2020328640/2020\|EPI_ISL_56965 | ···················· | ···················· | ····················· | ···················· | ···················· | ···················· |
| K | 3775 | hCoV-19/USA/WA-UW-24322/2020\|EPI_ISL_570600\|2020-0 | ···················· | ···················· | ····················· | ···················· | ···················· | ···················· |
| K | 3776 | hCoV-19/USA/WA-UW-24785/2020\|EPI_ISL_570601\|2020-0 | ···················· | ·A·················· | ····················· | ···················· | ···················· | ···················· |
| K | 3777 | hCoV-19/USA/WA-UW-24910/2020\|EPI_ISL_570602\|2020-0 | ···················· | ···················· | ····················· | ···················· | ···················· | ···················· |
| K | 3778 | hCoV-19/USA/WA-UW-24958/2020\|EPI_ISL_570603\|2020-0 | ···················· | ···················· | ····················· | ···················· | ···················· | ···················· |
| K | 3779 | hCoV-19/USA/WA-UW-26856/2020\|EPI_ISL_570604\|2020-0 | ···················· | ···················· | ····················· | ···················· | ···················· | ···················· |
| K | 3780 | hCoV-19/USA/WA-UW-22346/2020\|EPI_ISL_570605\|2020-0 | ···················· | ···················· | ····················· | ···················· | ···················· | ···················· |
| K | 3781 | hCoV-19/USA/WA-UW-22348/2020\|EPI_ISL_570606\|2020-0 | ···················· | ···················· | ····················· | ···················· | ···················· | ···················· |
| K | 3782 | hCoV-19/USA/WA-UW-22451/2020\|EPI_ISL_570607\|2020-0 | ···················· | ···················· | ····················· | ···················· | ···················· | ···················· |
| K | 3783 | hCoV-19/USA/WA-UW-24890/2020\|EPI_ISL_570608\|2020-0 | ···················· | ···················· | ····················· | ···················· | ···················· | ···················· |
| K | 3784 | hCoV-19/USA/WA-UW-22731/2020\|EPI_ISL_570609\|2020-0 | ········T··········· | ···················· | ····················· | ···················· | ···················· | ···················· |
| K | 3785 | hCoV-19/USA/WA-UW-22852/2020\|EPI_ISL_570610\|2020-0 | ···················· | ·A·················· | ····················· | ···················· | ···················· | ···················· |
| K | 3786 | hCoV-19/USA/WA-UW-24891/2020\|EPI_ISL_570611\|2020-0 | ···················· | ·A·················· | ····················· | ···················· | ···················· | ···················· |
| K | 3787 | hCoV-19/USA/CT-UW-2653/2020\|EPI_ISL_570612\|2020-03 | ···················· | ···················· | ····················· | ···················· | ···················· | ···················· |
| K | 3788 | hCoV-19/USA/OR-UW-22276/2020\|EPI_ISL_570613\|2020-0 | ···················· | ···················· | ····················· | ···················· | ···················· | ···················· |
| K | 3789 | hCoV-19/USA/OR-UW-22416/2020\|EPI_ISL_570614\|2020-0 | ···················· | ·A·················· | ····················· | ···················· | ···················· | ···················· |
| K | 3790 | hCoV-19/USA/OR-UW-22446/2020\|EPI_ISL_570615\|2020-0 | ···················· | ···················· | ····················· | ···················· | ···················· | ···················· |
| K | 3791 | hCoV-19/USA/OR-UW-22452/2020\|EPI_ISL_570616\|2020-0 | ···················· | ···················· | ····················· | ···················· | ···················· | ···················· |
| K | 3792 | hCoV-19/USA/OR-UW-22482/2020\|EPI_ISL_570617\|2020-0 | ···················· | ···················· | ····················· | ···················· | ···················· | ···················· |
| K | 3793 | hCoV-19/USA/OR-UW-22871/2020\|EPI_ISL_570618\|2020-0 | ···················· | ···················· | ····················· | ···················· | ···················· | ···················· |
| K | 3794 | hCoV-19/USA/OR-UW-22918/2020\|EPI_ISL_570619\|2020-0 | ···················· | ···················· | ····················· | ···················· | ···················· | ···················· |
| K | 3795 | hCoV-19/USA/WA-UW-22051/2020\|EPI_ISL_570620\|2020-0 | ···················· | ·A·················· | ····················· | ···················· | ···················· | ···················· |
| K | 3796 | hCoV-19/USA/WA-UW-22061/2020\|EPI_ISL_570621\|2020-0 | ···················· | ···················· | ····················· | ···················· | ···················· | ···················· |
| K | 3797 | hCoV-19/USA/WA-UW-22077/2020\|EPI_ISL_570622\|2020-0 | ···················· | ·A·················· | ····················· | ···················· | ···················· | ···················· |
| K | 3798 | hCoV-19/USA/WA-UW-22080/2020\|EPI_ISL_570623\|2020-0 | ···················· | ···················· | ····················· | ···················· | ···················· | ···················· |
| K | 3799 | hCoV-19/USA/WA-UW-22094/2020\|EPI_ISL_570624\|2020-0 | ···················· | ···················· | ····················· | ···················· | ···················· | ···················· |
| K | 3800 | hCoV-19/USA/WA-UW-22096/2020\|EPI_ISL_570625\|2020-0 | ···················· | ···················· | ····················· | ···················· | ···················· | ···················· |
| K | 3801 | hCoV-19/USA/WA-UW-22112/2020\|EPI_ISL_570626\|2020-0 | ···················· | ·A·················· | ····················· | ···················· | ···················· | ···················· |
| K | 3802 | hCoV-19/USA/WA-UW-22122/2020\|EPI_ISL_570627\|2020-0 | ···················· | ···················· | ····················· | ···················· | ···················· | ···················· |
| K | 3803 | hCoV-19/USA/WA-UW-22124/2020\|EPI_ISL_570628\|2020-0 | ···················· | ·A·················· | ····················· | ···················· | ···················· | ···················· |
| K | 3804 | hCoV-19/USA/WA-UW-22144/2020\|EPI_ISL_570629\|2020-0 | ···················· | ···················· | ····················· | ···················· | ···················· | ···················· |
| K | 3805 | hCoV-19/USA/WA-UW-22148/2020\|EPI_ISL_570630\|2020-0 | ···················· | ·A·················· | ····················· | ···················· | ···················· | ···················· |
| K | 3806 | hCoV-19/USA/WA-UW-22149/2020\|EPI_ISL_570631\|2020-0 | ···················· | ·A·················· | ····················· | ···················· | ···················· | ···················· |
| K | 3807 | hCoV-19/USA/WA-UW-22151/2020\|EPI_ISL_570632\|2020-0 | ···················· | ···················· | ····················· | ···················· | ···················· | ···················· |
| K | 3808 | hCoV-19/USA/WA-UW-22157/2020\|EPI_ISL_570633\|2020-0 | ···················· | ···················· | ····················· | ···················· | ···················· | ···················· |
| K | 3809 | hCoV-19/USA/WA-UW-22173/2020\|EPI_ISL_570634\|2020-0 | ···················· | ·A·················· | ····················· | ···················· | ···················· | ···················· |
| K | 3810 | hCoV-19/USA/WA-UW-22175/2020\|EPI_ISL_570635\|2020-0 | ···················· | ···················· | ····················· | ···················· | ···················· | ···················· |
| K | 3811 | hCoV-19/USA/WA-UW-22177/2020\|EPI_ISL_570636\|2020-0 | ···················· | ·A·················· | ····················· | ···················· | ···················· | ···················· |
| K | 3812 | hCoV-19/USA/WA-UW-22180/2020\|EPI_ISL_570637\|2020-0 | ···················· | ···················· | ····················· | ···················· | ···················· | ···················· |
| K | 3813 | hCoV-19/USA/WA-UW-22186/2020\|EPI_ISL_570638\|2020-0 | ···················· | ···················· | ····················· | ···················· | ···················· | ···················· |
| K | 3814 | hCoV-19/USA/WA-UW-22201/2020\|EPI_ISL_570639\|2020-0 | ···················· | ·A·················· | ····················· | ···················· | ···················· | ···················· |
| K | 3815 | hCoV-19/USA/WA-UW-22327/2020\|EPI_ISL_570660\|2020-0 | ···················· | ···················· | ····················· | ···················· | ···················· | ···················· |
| K | 3816 | hCoV-19/USA/WA-UW-22328/2020\|EPI_ISL_570661\|2020-0 | ···················· | ···················· | ····················· | ···················· | ···················· | ···················· |
| K | 3817 | hCoV-19/USA/WA-UW-22330/2020\|EPI_ISL_570662\|2020-0 | ···················· | ···················· | ····················· | ···················· | ···················· | ···················· |
| K | 3818 | hCoV-19/USA/WA-UW-22331/2020\|EPI_ISL_570663\|2020-0 | ···················· | ···················· | ····················· | ···················· | ···················· | ···················· |
| K | 3819 | hCoV-19/USA/WA-UW-22339/2020\|EPI_ISL_570664\|2020-0 | ···················· | ···················· | ····················· | ···················· | ···················· | ···················· |
| K | 3820 | hCoV-19/USA/WA-UW-22343/2020\|EPI_ISL_570665\|2020-0 | ···················· | ···················· | ····················· | ···················· | ···················· | ···················· |
| K | 3821 | hCoV-19/USA/WA-UW-22358/2020\|EPI_ISL_570666\|2020-0 | ···················· | ···················· | ····················· | ···················· | ···················· | ···················· |
| K | 3822 | hCoV-19/USA/WA-UW-22360/2020\|EPI_ISL_570667\|2020-0 | ···················· | ···················· | ····················· | ···················· | ···················· | ···················· |
| K | 3823 | hCoV-19/USA/WA-UW-22422/2020\|EPI_ISL_570682\|2020-0 | ···················· | ···················· | ····················· | ···················· | ···················· | ···················· |
| K | 3824 | hCoV-19/USA/WA-UW-22426/2020\|EPI_ISL_570683\|2020-0 | ···················· | ···················· | ····················· | ···················· | ···················· | ···················· |
| K | 3825 | hCoV-19/USA/WA-UW-22429/2020\|EPI_ISL_570684\|2020-0 | ···················· | ···················· | ····················· | ···················· | ···················· | ···················· |
| K | 3826 | hCoV-19/USA/WA-UW-22431/2020\|EPI_ISL_570685\|2020-0 | ···················· | ·A·················· | ····················· | ···················· | ···················· | ···················· |
| K | 3827 | hCoV-19/USA/WA-UW-22432/2020\|EPI_ISL_570686\|2020-0 | ···················· | ···················· | ····················· | ···················· | ···················· | ···················· |
| K | 3828 | hCoV-19/USA/WA-UW-22436/2020\|EPI_ISL_570687\|2020-0 | ···················· | ·A·················· | ····················· | ···················· | ···················· | ···················· |
| K | 3829 | hCoV-19/USA/WA-UW-22444/2020\|EPI_ISL_570688\|2020-0 | ···················· | ···················· | ····················· | ···················· | ···················· | ···················· |

| | | | | | | | | | |
|---|---|---|---|---|---|---|---|---|---|
| K | 3830 | hCoV-19/USA/WA-UW-22445/2020\|EPI_ISL_570689\|2020-0 | ···················· | ···················· | ····················· | ···················· | ···················· | ···················· | |
| K | 3831 | hCoV-19/USA/WA-UW-22447/2020\|EPI_ISL_570690\|2020-0 | ···················· | ·A·················· | ····················· | ···················· | ···················· | ···················· | |
| K | 3832 | hCoV-19/USA/WA-UW-22449/2020\|EPI_ISL_570691\|2020-0 | ···················· | ···················· | ····················· | ···················· | ···················· | ···················· | |
| K | 3833 | hCoV-19/USA/WA-UW-22454/2020\|EPI_ISL_570692\|2020-0 | ···················· | ·A·················· | ····················· | ···················· | ···················· | ···················· | |
| K | 3834 | hCoV-19/USA/WA-UW-22458/2020\|EPI_ISL_570693\|2020-0 | ···················· | ···················· | ····················· | ···················· | ···················· | ···················· | |
| K | 3835 | hCoV-19/USA/WA-UW-22468/2020\|EPI_ISL_570694\|2020-0 | ···················· | ·A·················· | ····················· | ···················· | ···················· | ···················· | |
| K | 3836 | hCoV-19/USA/WA-UW-22476/2020\|EPI_ISL_570695\|2020-0 | ···················· | ·A·················· | ····················· | ···················· | ···················· | ···················· | |
| K | 3837 | hCoV-19/USA/WA-UW-22491/2020\|EPI_ISL_570696\|2020-0 | ···················· | ···················· | ····················· | ···················· | ···················· | ···················· | |
| K | 3838 | hCoV-19/USA/WA-UW-22493/2020\|EPI_ISL_570697\|2020-0 | ···················· | ···················· | ····················· | ···················· | ···················· | ···················· | |
| K | 3839 | hCoV-19/USA/WA-UW-22495/2020\|EPI_ISL_570698\|2020-0 | ···················· | ·A·················· | ····················· | ···················· | ···················· | ···················· | |
| K | 3840 | hCoV-19/USA/WA-UW-22502/2020\|EPI_ISL_570699\|2020-0 | ···················· | ·A·················· | ····················· | ···················· | ···················· | ···················· | |
| K | 3842 | hCoV-19/Australia/VIC11976/2020\|EPI_ISL_563501\|202 | ···················· | ···················· | ····················· | ···················· | ···················· | ···················· | |
| K | 3843 | hCoV-19/Australia/VIC11978/2020\|EPI_ISL_563502\|202 | ···················· | ···················· | ····················· | ···················· | ···················· | ···················· | |
| K | 3844 | hCoV-19/Australia/VIC11996/2020\|EPI_ISL_563503\|202 | ···················· | ···················· | ····················· | ···················· | ···················· | ···················· | |
| K | 3845 | hCoV-19/Australia/VIC11997/2020\|EPI_ISL_563504\|202 | ···················· | ···················· | ····················· | ···················· | ···················· | ···················· | |
| K | 3846 | hCoV-19/Australia/VIC12010/2020\|EPI_ISL_563505\|202 | ···················· | ···················· | ····················· | ···················· | ···················· | ···················· | |
| K | 3847 | hCoV-19/Australia/VIC12014/2020\|EPI_ISL_563506\|202 | ···················· | ···················· | ····················· | ···················· | ···················· | ···················· | |
| K | 3848 | hCoV-19/Australia/VIC12018/2020\|EPI_ISL_563507\|202 | ···················· | ···················· | ····················· | ···················· | ···················· | ···················· | |
| K | 3849 | hCoV-19/Australia/VIC12020/2020\|EPI_ISL_563508\|202 | ···················· | ···················· | ····················· | ···················· | ···················· | ···················· | |
| K | 3850 | hCoV-19/Australia/VIC12031/2020\|EPI_ISL_563509\|202 | ···················· | ···················· | ····················· | ···················· | ···················· | ···················· | |
| K | 3851 | hCoV-19/Australia/VIC12034/2020\|EPI_ISL_563510\|202 | ···················· | ···················· | ····················· | ···················· | ···················· | ···················· | |
| K | 3852 | hCoV-19/Australia/VIC12036/2020\|EPI_ISL_563511\|202 | ···················· | ···················· | ····················· | ···················· | ···················· | ···················· | |
| K | 3853 | hCoV-19/Australia/VIC12037/2020\|EPI_ISL_563512\|202 | ···················· | ···················· | ····················· | ···················· | ···················· | ···················· | |
| K | 3854 | hCoV-19/Australia/VIC12039/2020\|EPI_ISL_563513\|202 | ···················· | ···················· | ····················· | ···················· | ···················· | ···················· | |
| K | 3855 | hCoV-19/Australia/VIC12061/2020\|EPI_ISL_563514\|202 | ···················· | ···················· | ····················· | ···················· | ···················· | ···················· | |
| K | 3856 | hCoV-19/Australia/VIC12063/2020\|EPI_ISL_563515\|202 | ···················· | ···················· | ····················· | ···················· | ···················· | ···················· | |
| K | 3857 | hCoV-19/Australia/VIC12066/2020\|EPI_ISL_563516\|202 | ···················· | ···················· | ····················· | ···················· | ···················· | ···················· | |
| K | 3858 | hCoV-19/Australia/VIC12077/2020\|EPI_ISL_563517\|202 | ···················· | ···················· | ····················· | ···················· | ···················· | ···················· | |
| K | 3859 | hCoV-19/Australia/VIC12081/2020\|EPI_ISL_563518\|202 | ···················· | ···················· | ····················· | ···················· | ···················· | ···················· | |
| K | 3860 | hCoV-19/Australia/VIC12084/2020\|EPI_ISL_563519\|202 | ···················· | ···················· | ····················· | ···················· | ···················· | ···················· | |
| K | 3861 | hCoV-19/Australia/VIC12086/2020\|EPI_ISL_563520\|202 | ···················· | ···················· | ····················· | ···················· | ···················· | ···················· | |
| K | 3862 | hCoV-19/Australia/VIC12088/2020\|EPI_ISL_563521\|202 | ···················· | ···················· | ····················· | ···················· | ···················· | ···················· | |
| K | 3863 | hCoV-19/Australia/VIC12100/2020\|EPI_ISL_563522\|202 | ···················· | ···················· | ····················· | ···················· | ···················· | ···················· | |
| K | 3864 | hCoV-19/Australia/VIC12104/2020\|EPI_ISL_563523\|202 | ···················· | ···················· | ····················· | ···················· | ···················· | ···················· | |
| K | 3867 | hCoV-19/Australia/VIC12126/2020\|EPI_ISL_563526\|202 | ···················· | ···················· | ····················· | ···················· | ···················· | ···················· | |
| K | 3870 | hCoV-19/Australia/VIC12143/2020\|EPI_ISL_563529\|202 | ···················· | ···················· | ····················· | ···················· | ···················· | ···················· | |
| K | 3871 | hCoV-19/Australia/VIC12157/2020\|EPI_ISL_563530\|202 | ···················· | ···················· | ····················· | ···················· | ···················· | ···················· | |
| K | 3872 | hCoV-19/Australia/VIC12163/2020\|EPI_ISL_563531\|202 | ···················· | ···················· | ····················· | ···················· | ···················· | ···················· | |
| K | 3873 | hCoV-19/Australia/VIC12168/2020\|EPI_ISL_563532\|202 | ···················· | ···················· | ····················· | ···················· | ···················· | ···················· | |
| K | 3874 | hCoV-19/Australia/VIC12169/2020\|EPI_ISL_563533\|202 | ···················· | ···················· | ····················· | ···················· | ···················· | ··················NN | 1 |
| K | 3875 | hCoV-19/Australia/VIC12177/2020\|EPI_ISL_563534\|202 | ···················· | ···················· | ····················· | ···················· | ···················· | ···················· | |
| K | 3876 | hCoV-19/Australia/VIC12181/2020\|EPI_ISL_563535\|202 | ···················· | ···················· | ····················· | ···················· | ···················· | ···················· | |
| K | 3877 | hCoV-19/Australia/VIC12183/2020\|EPI_ISL_563536\|202 | ···················· | ······A············· | ····················· | ···················· | ···················· | ···················· | |
| K | 3879 | hCoV-19/Australia/VIC12207/2020\|EPI_ISL_563538\|202 | ···················· | ···················· | ····················· | ···················· | ···················· | ···················· | |
| K | 3880 | hCoV-19/Australia/VIC12209/2020\|EPI_ISL_563539\|202 | ···················· | ···················· | ····················· | ···················· | ···················· | ···················· | |
| K | 3881 | hCoV-19/Australia/VIC12210/2020\|EPI_ISL_563540\|202 | ···················· | ···················· | ····················· | ···················· | ···················· | ···················· | |
| K | 3883 | hCoV-19/Australia/VIC12223/2020\|EPI_ISL_563542\|202 | ···················· | ···················· | ····················· | ···················· | ···················· | ···················· | |
| K | 3884 | hCoV-19/Australia/VIC12226/2020\|EPI_ISL_563543\|202 | ···················· | ···················· | ····················· | ···················· | ···················· | ···················· | |
| K | 3886 | hCoV-19/Australia/VIC12236/2020\|EPI_ISL_563545\|202 | ···················· | ···················· | ····················· | ···················· | ···················· | ···················· | |
| K | 3887 | hCoV-19/Australia/VIC12237/2020\|EPI_ISL_563546\|202 | ···················· | ···················· | ····················· | ···················· | ···················· | ···················· | |
| K | 3888 | hCoV-19/Australia/VIC12240/2020\|EPI_ISL_563547\|202 | ···················· | ···················· | ····················· | ···················· | ···················· | ···················· | |
| K | 3889 | hCoV-19/Australia/VIC12244/2020\|EPI_ISL_563548\|202 | ···················· | ···················· | ····················· | ···················· | ···················· | ···················· | |
| K | 3891 | hCoV-19/Australia/VIC12248/2020\|EPI_ISL_563550\|202 | ···················· | ···················· | ····················· | ···················· | ···················· | ···················· | |
| K | 3893 | hCoV-19/Australia/VIC12350/2020\|EPI_ISL_563552\|202 | ···················· | ···················· | ····················· | ···················· | ···················· | ···················· | |
| K | 3894 | hCoV-19/Australia/VIC12352/2020\|EPI_ISL_563553\|202 | ···················· | ···················· | ····················· | ···················· | ···················· | ···················· | |
| K | 3895 | hCoV-19/Australia/VIC12604/2020\|EPI_ISL_563576\|202 | ···················· | ···················· | ····················· | ···················· | ···················· | ···················· | |
| K | 3896 | hCoV-19/Australia/VIC12661/2020\|EPI_ISL_563577\|202 | ···················· | ···················· | ····················· | ···················· | ···················· | ···················· | |
| K | 3897 | hCoV-19/Australia/VIC12694/2020\|EPI_ISL_563578\|202 | ···················· | ···················· | ····················· | ···················· | ···················· | ···················· | |
| K | 3898 | hCoV-19/Australia/VIC12696/2020\|EPI_ISL_563579\|202 | ···················· | ···················· | ····················· | ···················· | ···················· | ···················· | |
| K | 3899 | hCoV-19/Australia/VIC12710/2020\|EPI_ISL_563580\|202 | ···················· | ···················· | ····················· | ···················· | ···················· | ···················· | |
| K | 3900 | hCoV-19/Australia/VIC12712/2020\|EPI_ISL_563581\|202 | ···················· | ···················· | ····················· | ···················· | ···················· | ···················· | |
| K | 3901 | hCoV-19/Australia/VIC12715/2020\|EPI_ISL_563582\|202 | ···················· | ···················· | ····················· | ···················· | ···················· | ···················· | |
| K | 3902 | hCoV-19/Australia/VIC12733/2020\|EPI_ISL_563583\|202 | ···················· | ···················· | ····················· | ···················· | ···················· | ···················· | |
| K | 3903 | hCoV-19/Australia/VIC12734/2020\|EPI_ISL_563584\|202 | ···················· | ···················· | ····················· | ···················· | ···················· | ···················· | |
| K | 3904 | hCoV-19/Australia/VIC12737/2020\|EPI_ISL_563585\|202 | ···················· | ···················· | ····················· | ···················· | ···················· | ···················· | |
| K | 3905 | hCoV-19/Australia/VIC12742/2020\|EPI_ISL_563586\|202 | ···················· | ···················· | ····················· | ···················· | ···················· | ···················· | |
| K | 3906 | hCoV-19/Australia/VIC12770/2020\|EPI_ISL_563590\|202 | ···················· | ···················· | ····················· | ···················· | ···················· | ···················· | |
| K | 3908 | hCoV-19/Australia/VIC12773/2020\|EPI_ISL_563592\|202 | ···················· | ···················· | ····················· | ···················· | ···················· | ···················· | |
| K | 3909 | hCoV-19/Australia/VIC12776/2020\|EPI_ISL_563593\|202 | ···················· | ···················· | ····················· | ···················· | ···················· | ···················· | |
| K | 3910 | hCoV-19/Australia/VIC12803/2020\|EPI_ISL_563596\|202 | ···················· | ···················· | ····················· | ···················· | ···················· | ···················· | |
| K | 3911 | hCoV-19/Australia/VIC12809/2020\|EPI_ISL_563597\|202 | ···················· | ···················· | ····················· | ···················· | ···················· | ···················· | |
| K | 3913 | hCoV-19/Australia/VIC12833/2020\|EPI_ISL_563599\|202 | ···················· | ···················· | ····················· | ···················· | ···················· | ···················· | |
| K | 3914 | hCoV-19/Australia/VIC8008/2020\|EPI_ISL_564800\|2020 | ···················· | ···················· | ····················· | ···················· | ···················· | ···················· | |
| K | 3915 | hCoV-19/Australia/VIC8009/2020\|EPI_ISL_564801\|2020 | ···················· | ···················· | ····················· | ···················· | ···················· | ···················· | |
| K | 3916 | hCoV-19/Australia/VIC8012/2020\|EPI_ISL_564802\|2020 | ···················· | ···················· | ····················· | ···················· | ···················· | ···················· | |
| K | 3917 | hCoV-19/Australia/VIC8014/2020\|EPI_ISL_564803\|2020 | ···················· | ···················· | ····················· | ···················· | ···················· | ···················· | |
| K | 3918 | hCoV-19/Australia/VIC8016/2020\|EPI_ISL_564804\|2020 | ···················· | ···················· | ····················· | ···················· | ···················· | ···················· | |
| K | 3919 | hCoV-19/Australia/VIC8018/2020\|EPI_ISL_564805\|2020 | ···················· | ···················· | ····················· | ···················· | ···················· | ···················· | |
| K | 3920 | hCoV-19/Australia/VIC8020/2020\|EPI_ISL_564806\|2020 | ···················· | ···················· | ····················· | ···················· | ···················· | ···················· | |
| K | 3921 | hCoV-19/Australia/VIC8022/2020\|EPI_ISL_564807\|2020 | ···················· | ···················· | ····················· | ···················· | ···················· | ···················· | |

K 3922 hCoV-19/Australia/VIC8023/2020|EPI_ISL_564808|2020 ···················· ···················· ····················· ···················· ···················· ····················
K 3923 hCoV-19/Australia/VIC8027/2020|EPI_ISL_564809|2020 ···················· ···················· ····················· ···················· ···················· ····················
K 3924 hCoV-19/Australia/VIC8028/2020|EPI_ISL_564810|2020 ···················· ···················· ····················· ···················· ···················· ····················
K 3925 hCoV-19/Australia/VIC8030/2020|EPI_ISL_564811|2020 ···················· ···················· ····················· ···················· ···················· ····················
K 3926 hCoV-19/Australia/VIC8032/2020|EPI_ISL_564812|2020 ···················· ···················· ····················· ···················· ···················· ····················
K 3927 hCoV-19/Australia/VIC8033/2020|EPI_ISL_564813|2020 ···················· ···················· ····················· ···················· ···················· ····················
K 3928 hCoV-19/Australia/VIC8037/2020|EPI_ISL_564814|2020 ···················· ···················· ····················· ···················· ···················· ····················
K 3929 hCoV-19/Australia/VIC8040/2020|EPI_ISL_564815|2020 ···················· ···················· ····················· ···················· ···················· ····················
K 3930 hCoV-19/Australia/VIC8046/2020|EPI_ISL_564816|2020 ···················· ···················· ····················· ···················· ···················· ····················
K 3931 hCoV-19/Australia/VIC8047/2020|EPI_ISL_564817|2020 ···················· ···················· ····················· ···················· ···················· ····················
K 3932 hCoV-19/Australia/VIC8050/2020|EPI_ISL_564818|2020 ···················· ···················· ····················· ···················· ···················· ····················
K 3933 hCoV-19/Australia/VIC8051/2020|EPI_ISL_564819|2020 ···················· ···················· ····················· ···················· ···················· ····················
K 3934 hCoV-19/Australia/VIC8052/2020|EPI_ISL_564820|2020 ···················· ···················· ····················· ···················· ···················· ····················
K 3935 hCoV-19/Australia/VIC8053/2020|EPI_ISL_564821|2020 ···················· ···················· ····················· ···················· ···················· ····················
K 3936 hCoV-19/Australia/VIC8055/2020|EPI_ISL_564822|2020 ···················· ···················· ····················· ···················· ···················· ····················
K 3937 hCoV-19/Australia/VIC8058/2020|EPI_ISL_564823|2020 ···················· ···················· ····················· ···················· ···················· ····················
K 3938 hCoV-19/Australia/VIC8059/2020|EPI_ISL_564824|2020 ···················· ···················· ····················· ···················· ···················· ····················
K 3939 hCoV-19/Australia/VIC8060/2020|EPI_ISL_564825|2020 ···················· ···················· ····················· ···················· ···················· ····················
K 3940 hCoV-19/Australia/VIC8061/2020|EPI_ISL_564826|2020 ···················· ···················· ····················· ···················· ···················· ····················
K 3941 hCoV-19/Australia/VIC8062/2020|EPI_ISL_564827|2020 ···················· ···················· ····················· ···················· ···················· ····················
K 3942 hCoV-19/Australia/VIC8063/2020|EPI_ISL_564828|2020 ···················· ···················· ····················· ···················· ···················· ····················
K 3943 hCoV-19/Australia/VIC8067/2020|EPI_ISL_564829|2020 ···················· ···················· ····················· ···················· ···················· ····················
K 3944 hCoV-19/Australia/VIC8069/2020|EPI_ISL_564830|2020 ···················· ···················· ····················· ···················· ···················· ····················
K 3945 hCoV-19/Australia/VIC8070/2020|EPI_ISL_564831|2020 ···················· ···················· ····················· ···················· ···················· ····················
K 3946 hCoV-19/Australia/VIC8073/2020|EPI_ISL_564832|2020 ···················· ···················· ····················· ···················· ···················· ····················
K 3947 hCoV-19/Australia/VIC8074/2020|EPI_ISL_564833|2020 ···················· ···················· ····················· ···················· ···················· ····················
K 3948 hCoV-19/Australia/VIC8075/2020|EPI_ISL_564834|2020 ···················· ···················· ····················· ···················· ···················· ····················
K 3949 hCoV-19/Australia/VIC8077/2020|EPI_ISL_564835|2020 ···················· ···················· ····················· ···················· ···················· ····················
K 3950 hCoV-19/Australia/VIC8078/2020|EPI_ISL_564836|2020 ···················· ···················· ····················· ···················· ···················· ····················
K 3951 hCoV-19/Australia/VIC8081/2020|EPI_ISL_564837|2020 ···················· ···················· ····················· ···················· ···················· ····················
K 3952 hCoV-19/Australia/VIC8082/2020|EPI_ISL_564838|2020 ···················· ···················· ····················· ···················· ···················· ····················
K 3953 hCoV-19/Australia/VIC8085/2020|EPI_ISL_564839|2020 ···················· ···················· ····················· ···················· ···················· ····················
K 3954 hCoV-19/Australia/VIC8086/2020|EPI_ISL_564840|2020 ···················· ···················· ····················· ···················· ···················· ····················
K 3955 hCoV-19/Australia/VIC8087/2020|EPI_ISL_564841|2020 ···················· ···················· ····················· ···················· ···················· ····················
K 3956 hCoV-19/Australia/VIC8091/2020|EPI_ISL_564842|2020 ···················· ···················· ····················· ···················· ···················· ····················
K 3957 hCoV-19/Australia/VIC8092/2020|EPI_ISL_564843|2020 ···················· ···················· ····················· ···················· ···················· ····················
K 3958 hCoV-19/Australia/VIC8094/2020|EPI_ISL_564844|2020 ···················· ···················· ····················· ···················· ···················· ····················
K 3959 hCoV-19/Australia/VIC8096/2020|EPI_ISL_564845|2020 ···················· ···················· ····················· ···················· ···················· ····················
K 3960 hCoV-19/Australia/VIC8098/2020|EPI_ISL_564846|2020 ···················· ···················· ····················· ···················· ···················· ····················
K 3961 hCoV-19/Australia/VIC8099/2020|EPI_ISL_564847|2020 ···················· ···················· ····················· ···················· ···················· ····················
K 3963 hCoV-19/Australia/VIC8102/2020|EPI_ISL_564849|2020 ···················· ···················· ····················· ···················· ···················· ····················
K 3964 hCoV-19/Australia/VIC8103/2020|EPI_ISL_564850|2020 ···················· ···················· ····················· ···················· ···················· ····················
K 3965 hCoV-19/Australia/VIC8104/2020|EPI_ISL_564851|2020 ···················· ···················· ····················· ···················· ···················· ····················
K 3966 hCoV-19/Australia/VIC8105/2020|EPI_ISL_564852|2020 ···················· ···················· ····················· ···················· ···················· ····················
K 3967 hCoV-19/Australia/VIC8107/2020|EPI_ISL_564853|2020 ···················· ···················· ····················· ···················· ···················· ····················
K 3968 hCoV-19/Australia/VIC8108/2020|EPI_ISL_564854|2020 ···················· ···················· ····················· ···················· ···················· ····················
K 3969 hCoV-19/Australia/VIC8110/2020|EPI_ISL_564855|2020 ···················· ···················· ····················· ···················· ···················· ····················
K 3970 hCoV-19/Australia/VIC8113/2020|EPI_ISL_564856|2020 ···················· ···················· ····················· ···················· ···················· ····················
K 3971 hCoV-19/Australia/VIC8114/2020|EPI_ISL_564857|2020 ···················· ···················· ····················· ···················· ···················· ····················
K 3972 hCoV-19/Australia/VIC8115/2020|EPI_ISL_564858|2020 ···················· ···················· ····················· ···················· ···················· ····················
K 3973 hCoV-19/Australia/VIC8120/2020|EPI_ISL_564859|2020 ···················· ···················· ····················· ···················· ···················· ····················
K 3974 hCoV-19/Australia/VIC8121/2020|EPI_ISL_564860|2020 ···················· ···················· ····················· ···················· ···················· ····················
K 3975 hCoV-19/Australia/VIC8122/2020|EPI_ISL_564861|2020 ···················· ···················· ····················· ···················· ···················· ····················
K 3976 hCoV-19/Australia/VIC8123/2020|EPI_ISL_564862|2020 ···················· ···················· ····················· ···················· ···················· ····················
K 3977 hCoV-19/Australia/VIC8124/2020|EPI_ISL_564863|2020 ···················· ···················· ····················· ···················· ···················· ····················
K 3978 hCoV-19/Australia/VIC8125/2020|EPI_ISL_564864|2020 ···················· ···················· ····················· ···················· ···················· ····················
K 3979 hCoV-19/Australia/VIC8127/2020|EPI_ISL_564865|2020 ···················· ···················· ····················· ···················· ···················· ····················
K 3980 hCoV-19/Australia/VIC8129/2020|EPI_ISL_564866|2020 ···················· ···················· ····················· ···················· ···················· ····················
K 3981 hCoV-19/Australia/VIC8130/2020|EPI_ISL_564867|2020 ···················· ···················· ····················· ···················· ···················· ····················
K 3982 hCoV-19/Australia/VIC8131/2020|EPI_ISL_564868|2020 ···················· ···················· ····················· ···················· ···················· ····················
K 3983 hCoV-19/Australia/VIC8132/2020|EPI_ISL_564869|2020 ···················· ···················· ····················· ···················· ···················· ····················
K 3984 hCoV-19/Australia/VIC8138/2020|EPI_ISL_564870|2020 ···················· ···················· ····················· ···················· ···················· ····················
K 3985 hCoV-19/Australia/VIC8143/2020|EPI_ISL_564871|2020 ·····G·············· ···················· ····················· ···················· ···················· ····················
K 3986 hCoV-19/Australia/VIC8145/2020|EPI_ISL_564872|2020 ···················· ···················· ····················· ···················· ···················· ····················
K 3987 hCoV-19/Australia/VIC8147/2020|EPI_ISL_564873|2020 ···················· ···················· ····················· ···················· ···················· ····················
K 3988 hCoV-19/Australia/VIC8150/2020|EPI_ISL_564874|2020 ···················· ···················· ····················· ···················· ···················· ····················
K 3989 hCoV-19/Australia/VIC8153/2020|EPI_ISL_564875|2020 ···················· ···················· ····················· ···················· ···················· ····················
K 3990 hCoV-19/Australia/VIC8157/2020|EPI_ISL_564876|2020 ···················· ···················· ····················· ···················· ···················· ····················
K 3991 hCoV-19/Australia/VIC8159/2020|EPI_ISL_564877|2020 ···················· ···················· ····················· ···················· ···················· ····················
K 3992 hCoV-19/Australia/VIC8160/2020|EPI_ISL_564878|2020 ···················· ···················· ····················· ···················· ···················· ····················
K 3993 hCoV-19/Australia/VIC8161/2020|EPI_ISL_564879|2020 ···················· ···················· ····················· ···················· ···················· ····················
K 3994 hCoV-19/Australia/VIC8164/2020|EPI_ISL_564880|2020 ···················· ···················· ····················· ···················· ···················· ····················
K 3995 hCoV-19/Australia/VIC8165/2020|EPI_ISL_564881|2020 ···················· ···················· ····················· ···················· ···················· ····················
K 3996 hCoV-19/Australia/VIC8169/2020|EPI_ISL_564882|2020 ···················· ···················· ····················· ···················· ···················· ····················
K 3997 hCoV-19/Australia/VIC8171/2020|EPI_ISL_564883|2020 ···················· ···················· ····················· ···················· ···················· ····················
K 3998 hCoV-19/Australia/VIC8174/2020|EPI_ISL_564884|2020 ···················· ···················· ····················· ···················· ···················· ····················
K 3999 hCoV-19/England/QEUH-9C81BD/2020|EPI_ISL_567080|20 ···················· ···················· ····················· ···················· ······T············· ····················
K 4000 hCoV-19/England/QEUH-9C8A2B/2020|EPI_ISL_567081|20 ···················· ·A·················· ····················· ···················· ···················· ···A················
K 4001 hCoV-19/England/QEUH-9C9125/2020|EPI_ISL_567082|20 ···················· ···················· ····················· ···················· ···················· ····················
K 4002 hCoV-19/England/QEUH-9C8108/2020|EPI_ISL_567083|20 ···················· ···················· ····················· ···················· ···················· ····················

| | | | | | | | | |
|---|---|---|---|---|---|---|---|---|
| K | 4003 | hCoV-19/England/QEUH-9C7B92/2020\|EPI_ISL_567085\|20 | .................... | .A.................. | .................... | .................... | .................... | .................... |
| K | 4004 | hCoV-19/England/QEUH-9C7C53/2020\|EPI_ISL_567086\|20 | .................... | .................... | .................... | .................... | .................... | .................... |
| K | 4005 | hCoV-19/England/QEUH-9C8C34/2020\|EPI_ISL_567087\|20 | .................... | .................... | .................... | .................... | .................... | .................... |
| K | 4006 | hCoV-19/England/QEUH-9C8F2C/2020\|EPI_ISL_567088\|20 | .................... | .A.................. | .................... | .................... | .................... | .................... |
| K | 4007 | hCoV-19/England/QEUH-9C8663/2020\|EPI_ISL_567090\|20 | .................... | .................... | .................... | .................... | .................... | .................... |
| K | 4008 | hCoV-19/England/QEUH-9C8539/2020\|EPI_ISL_567091\|20 | .................... | .................... | .................... | .................... | .................... | .................... |
| K | 4009 | hCoV-19/England/QEUH-9C8B28/2020\|EPI_ISL_567093\|20 | .................... | .................... | .................... | .................... | .................... | .................... |
| K | 4010 | hCoV-19/England/QEUH-9C8609/2020\|EPI_ISL_567094\|20 | .................... | .................... | .................... | .................... | .................... | .................... |
| K | 4011 | hCoV-19/England/QEUH-9C89A6/2020\|EPI_ISL_567095\|20 | .................... | .................... | .................... | .................... | .................... | .................... |
| K | 4012 | hCoV-19/England/QEUH-9C8FD1/2020\|EPI_ISL_567096\|20 | .................... | .................... | .................... | .................... | .................... | .................... |
| K | 4013 | hCoV-19/England/QEUH-9C8AC1/2020\|EPI_ISL_567097\|20 | .................... | .................... | .................... | .................... | .................... | .................... |
| K | 4014 | hCoV-19/England/QEUH-9C8205/2020\|EPI_ISL_567098\|20 | .................... | .................... | .................... | .................... | .................... | .................... |
| K | 4015 | hCoV-19/England/QEUH-9C91DA/2020\|EPI_ISL_567099\|20 | .................... | .................... | .................... | .................... | .................... | .................... |
| K | 4016 | hCoV-19/England/ALDP-9BC90F/2020\|EPI_ISL_568207\|20 | .................... | .................... | .................... | .................... | .................... | .................... |
| K | 4017 | hCoV-19/England/ALDP-9BC83F/2020\|EPI_ISL_568208\|20 | .................... | .................... | .................... | .................... | .................... | .................... |
| K | 4018 | hCoV-19/England/ALDP-9BC741/2020\|EPI_ISL_568211\|20 | .................... | .A.................. | .................... | .................... | .................... | .................... |
| K | 4019 | hCoV-19/England/ALDP-9BEA64/2020\|EPI_ISL_568212\|20 | .................... | .................... | .................... | .................... | .................... | .................... |
| K | 4020 | hCoV-19/England/ALDP-9BE32D/2020\|EPI_ISL_568213\|20 | .................... | .................... | .................... | .................... | .................... | .................... |
| K | 4021 | hCoV-19/England/ALDP-9BDEE1/2020\|EPI_ISL_568214\|20 | .................... | .................... | .................... | .................... | .................... | .................... |
| K | 4022 | hCoV-19/England/ALDP-9BE518/2020\|EPI_ISL_568215\|20 | .................... | .................... | .................... | .................... | .................... | .................... |
| K | 4023 | hCoV-19/England/ALDP-9BE3C3/2020\|EPI_ISL_568216\|20 | .................... | .................... | .................... | .................... | .................... | .................... |
| K | 4024 | hCoV-19/England/ALDP-9BE2C6/2020\|EPI_ISL_568217\|20 | .................... | .................... | .................... | .................... | .................... | .................... |
| K | 4025 | hCoV-19/England/ALDP-9BDC7E/2020\|EPI_ISL_568218\|20 | .................... | .A.................. | .................... | .................... | .................... | .................... |
| K | 4026 | hCoV-19/England/ALDP-9BC77E/2020\|EPI_ISL_568219\|20 | .................... | .................... | .................... | .................... | .................... | .................... |
| K | 4027 | hCoV-19/England/ALDP-9BDF2A/2020\|EPI_ISL_568220\|20 | .................... | .A.................. | .................... | .................... | .................... | .................... |
| K | 4028 | hCoV-19/England/ALDP-9BE16F/2020\|EPI_ISL_568221\|20 | .................... | .................... | .................... | .................... | .................... | .................... |
| K | 4029 | hCoV-19/England/ALDP-9BC6DB/2020\|EPI_ISL_568222\|20 | .................... | .................... | .................... | .................... | .................... | .................... |
| K | 4030 | hCoV-19/Scotland/QEUH-9B82EA/2020\|EPI_ISL_568223\|2 | .................... | .................... | .................... | .................... | .................... | .................... |
| K | 4031 | hCoV-19/Scotland/QEUH-9B85F0/2020\|EPI_ISL_568224\|2 | .................... | .................... | .................... | .................... | .................... | .................... |
| K | 4032 | hCoV-19/Scotland/QEUH-9B8930/2020\|EPI_ISL_568225\|2 | .................... | .................... | .................... | .................... | .................... | .................... |
| K | 4033 | hCoV-19/Scotland/QEUH-9B86DF/2020\|EPI_ISL_568226\|2 | .................... | .................... | .................... | .................... | .................... | .................... |
| K | 4034 | hCoV-19/Scotland/QEUH-9B8657/2020\|EPI_ISL_568227\|2 | .................... | .................... | .................... | .................... | .................... | .................... |
| K | 4035 | hCoV-19/Scotland/QEUH-9B8D61/2020\|EPI_ISL_568228\|2 | .................... | .................... | .................... | .................... | .................... | .................... |
| K | 4036 | hCoV-19/Scotland/QEUH-9B8A4C/2020\|EPI_ISL_568241\|2 | .................... | .................... | .................... | .................... | .................... | .................... |
| K | 4037 | hCoV-19/Scotland/QEUH-9B8226/2020\|EPI_ISL_568242\|2 | .................... | .................... | .................... | .................... | .................... | .................... |
| K | 4038 | hCoV-19/Scotland/QEUH-9B9B93/2020\|EPI_ISL_568243\|2 | .................... | .................... | .................... | .................... | .................... | .................... |
| K | 4039 | hCoV-19/Scotland/QEUH-9B8921/2020\|EPI_ISL_568244\|2 | .................... | .................... | .................... | .................... | .................... | .................... |
| K | 4040 | hCoV-19/Scotland/QEUH-9B9A96/2020\|EPI_ISL_568245\|2 | .................... | .................... | .................... | .................... | .................... | .................... |
| K | 4041 | hCoV-19/Scotland/QEUH-9B8709/2020\|EPI_ISL_568246\|2 | .................... | .................... | .................... | .................... | .................... | .................... |
| K | 4042 | hCoV-19/Scotland/QEUH-9B86B1/2020\|EPI_ISL_568247\|2 | .................... | .................... | .................... | .................... | .................... | .................... |
| K | 4043 | hCoV-19/Scotland/QEUH-9B8912/2020\|EPI_ISL_568248\|2 | .................... | .................... | .................... | .................... | .................... | .................... |
| K | 4044 | hCoV-19/Scotland/QEUH-9B82CC/2020\|EPI_ISL_568249\|2 | .................... | .................... | .................... | .................... | .................... | .................... |
| K | 4045 | hCoV-19/France/PAC-IHU-0475/2020\|EPI_ISL_569538\|20 | .................... | .................... | .................... | .................... | .................... | .................... |
| K | 4046 | hCoV-19/France/PAC-IHU-0481/2020\|EPI_ISL_569540\|20 | .................... | .................... | .................... | .................... | .................... | .................... |
| K | 4047 | hCoV-19/France/PAC-IHU-0483/2020\|EPI_ISL_569541\|20 | .................... | .................... | .................... | .................... | .................... | .................... |
| K | 4048 | hCoV-19/France/PAC-IHU-0485/2020\|EPI_ISL_569542\|20 | .................... | .................... | .................... | .................... | .................... | .................... |
| K | 4049 | hCoV-19/France/PAC-IHU-0487/2020\|EPI_ISL_569543\|20 | .................... | .................... | .................... | .................... | .................... | .................... |
| K | 4050 | hCoV-19/France/PAC-IHU-0488/2020\|EPI_ISL_569544\|20 | .................... | .................... | .................... | .................... | .................... | .................... |
| K | 4051 | hCoV-19/France/PAC-IHU-0489/2020\|EPI_ISL_569545\|20 | .................... | .................... | .................... | .................... | .................... | .................... |
| K | 4052 | hCoV-19/France/PAC-IHU-0490/2020\|EPI_ISL_569546\|20 | .................... | .................... | .................... | .................... | .................... | .................... |
| K | 4053 | hCoV-19/France/PAC-IHU-0492/2020\|EPI_ISL_569547\|20 | .................... | .................... | .................... | .................... | .................... | .................... |
| K | 4054 | hCoV-19/France/PAC-IHU-0493/2020\|EPI_ISL_569548\|20 | .................... | .................... | .................... | .................... | .................... | .................... |
| K | 4055 | hCoV-19/France/PAC-IHU-0494/2020\|EPI_ISL_569549\|20 | .................... | .................... | .................... | .................... | .................... | .................... |
| K | 4056 | hCoV-19/France/PAC-IHU-0495/2020\|EPI_ISL_569550\|20 | .................... | .................... | .................... | .................... | .................... | .................... |
| K | 4057 | hCoV-19/France/PAC-IHU-0496/2020\|EPI_ISL_569551\|20 | .................... | .................... | .................... | .................... | .................... | .................... |
| K | 4058 | hCoV-19/France/PAC-IHU-0497/2020\|EPI_ISL_569552\|20 | .................... | .................... | .................... | .................... | .................... | .................... |
| K | 4059 | hCoV-19/France/PAC-IHU-0498/2020\|EPI_ISL_569553\|20 | .................... | .................... | .................... | .................... | .................... | .................... |
| K | 4060 | hCoV-19/France/PAC-IHU-0499/2020\|EPI_ISL_569554\|20 | .................... | .................... | .................... | .................... | .................... | .................... |
| K | 4061 | hCoV-19/France/PAC-IHU-0500/2020\|EPI_ISL_569555\|20 | .................... | .................... | .................... | .................... | .................... | .................... |
| K | 4062 | hCoV-19/France/PAC-IHU-0501/2020\|EPI_ISL_569556\|20 | .................... | .................... | .................... | .................... | .................... | .................... |
| K | 4063 | hCoV-19/France/PAC-IHU-0502/2020\|EPI_ISL_569557\|20 | .................... | .................... | .................... | .................... | .................... | .................... |
| K | 4064 | hCoV-19/France/PAC-IHU-0504/2020\|EPI_ISL_569558\|20 | .................... | .................... | .................... | .................... | .................... | .................... |
| K | 4065 | hCoV-19/France/PAC-IHU-0505/2020\|EPI_ISL_569559\|20 | .................... | .................... | .................... | .................... | .................... | .................... |
| K | 4066 | hCoV-19/France/PAC-IHU-0522/2020\|EPI_ISL_569570\|20 | .................... | .................... | .................... | .................... | .................... | .................... |
| K | 4067 | hCoV-19/France/PAC-IHU-0526/2020\|EPI_ISL_569572\|20 | .................... | .................... | .................... | .................... | .................... | .................... |
| K | 4068 | hCoV-19/France/PAC-IHU-0527/2020\|EPI_ISL_569573\|20 | .................... | .................... | .................... | .................... | .................... | .................... |
| K | 4069 | hCoV-19/France/PAC-IHU-0530/2020\|EPI_ISL_569574\|20 | .................... | .................... | .................... | .................... | .................... | .................... |
| K | 4070 | hCoV-19/France/PAC-IHU-0531/2020\|EPI_ISL_569575\|20 | .................... | .................... | .................... | .................... | .................... | .................... |
| K | 4071 | hCoV-19/France/PAC-IHU-0532/2020\|EPI_ISL_569576\|20 | .................... | .................... | .................... | .................... | .................... | .................... |
| K | 4072 | hCoV-19/France/PAC-IHU-0534/2020\|EPI_ISL_569577\|20 | .................... | .................... | .................... | .................... | .................... | .................... |
| K | 4073 | hCoV-19/France/PAC-IHU-0535/2020\|EPI_ISL_569578\|20 | .................... | .................... | .................... | .................... | .................... | .................... |
| K | 4074 | hCoV-19/USA/WA-UW-12377/2020\|EPI_ISL_570520\|2020-0 | .................... | .................... | .................... | .................... | .................... | .................... |
| K | 4075 | hCoV-19/USA/WA-UW-12378/2020\|EPI_ISL_570521\|2020-0 | .................... | .................... | .................... | .................... | .................... | .................... |
| K | 4076 | hCoV-19/USA/WA-UW-12384/2020\|EPI_ISL_570522\|2020-0 | .................... | .................... | .................... | .................... | .................... | .................... |
| K | 4077 | hCoV-19/USA/WA-UW-12388/2020\|EPI_ISL_570523\|2020-0 | .................... | .A.................. | .................... | .................... | .................... | .................... |
| K | 4078 | hCoV-19/USA/WA-UW-12389/2020\|EPI_ISL_570524\|2020-0 | .................... | .A.................. | .................... | .................... | .................... | .................... |
| K | 4079 | hCoV-19/USA/WA-UW-12390/2020\|EPI_ISL_570525\|2020-0 | .................... | .................... | .................... | .................... | .................... | .................... |
| K | 4080 | hCoV-19/USA/WA-UW-12391/2020\|EPI_ISL_570526\|2020-0 | ...........T........ | .................... | .................... | .................... | .................... | .................... |
| K | 4081 | hCoV-19/USA/WA-UW-12392/2020\|EPI_ISL_570527\|2020-0 | .................... | .................... | .................... | .................... | .................... | .................... |
| K | 4082 | hCoV-19/USA/WA-UW-12395/2020\|EPI_ISL_570528\|2020-0 | .................... | .................... | .................... | .................... | .................... | .................... |

| | | | | | | | | | |
|---|---|---|---|---|---|---|---|---|---|
| K | 4083 | hCoV-19/USA/WA-UW-12398/2020\|EPI_ISL_570529\|2020-0 | ···················· | ···················· | ···················· | ···················· | ···················· | ···················· | |
| K | 4084 | hCoV-19/USA/WA-UW-12400/2020\|EPI_ISL_570530\|2020-0 | ···················· | ···················· | ···················· | ···················· | ···················· | ···················· | |
| K | 4085 | hCoV-19/USA/WA-UW-12401/2020\|EPI_ISL_570531\|2020-0 | ···················· | ···················· | ···················· | ···················· | ···················· | ···················· | |
| K | 4086 | hCoV-19/USA/WA-UW-12436/2020\|EPI_ISL_570532\|2020-0 | ···················· | ···················· | ···················· | ···················· | ···················· | ···················· | |
| K | 4087 | hCoV-19/USA/WA-UW-12437/2020\|EPI_ISL_570533\|2020-0 | ···················· | ···················· | ···················· | ···················· | ···················· | ···················· | |
| K | 4088 | hCoV-19/USA/WA-UW-12440/2020\|EPI_ISL_570534\|2020-0 | ···················· | ···················· | ···················· | ···················· | ···················· | ···················· | |
| K | 4089 | hCoV-19/USA/WA-UW-12443/2020\|EPI_ISL_570535\|2020-0 | ···················· | ···················· | ···················· | ···················· | ···················· | ···················· | |
| K | 4090 | hCoV-19/USA/WA-UW-1245/2020\|EPI_ISL_570536\|2020-03 | ···················· | ···················· | ···················· | ···················· | ···················· | ···················· | |
| K | 4091 | hCoV-19/USA/WA-UW-1247/2020\|EPI_ISL_570537\|2020-03 | ···················· | ···················· | ···················· | ···················· | ···················· | ···················· | |
| K | 4092 | hCoV-19/USA/WA-UW-12478/2020\|EPI_ISL_570538\|2020-0 | ···················· | ···················· | ···················· | ···················· | ···················· | ···················· | |
| K | 4093 | hCoV-19/USA/WA-UW-12488/2020\|EPI_ISL_570539\|2020-0 | ···················· | ·A·················· | ···················· | ···················· | ···················· | ···················· | |
| K | 4094 | hCoV-19/USA/WA-UW-22048/2020\|EPI_ISL_570558\|2020-0 | ···················· | ···················· | ···················· | ···················· | ···················· | ···················· | |
| K | 4095 | hCoV-19/USA/WA-UW-22090/2020\|EPI_ISL_570559\|2020-0 | ···················· | ···················· | ···················· | ···················· | ···················· | ···················· | |
| K | 4096 | hCoV-19/USA/WA-UW-22117/2020\|EPI_ISL_570560\|2020-0 | ···················· | ·A·················· | ···················· | ···················· | ···················· | ···················· | |
| K | 4098 | hCoV-19/Australia/VIC11615/2020\|EPI_ISL_563434\|202 | ···················· | ···················· | ···················· | ···················· | ···················· | ···················· | |
| K | 4101 | hCoV-19/Australia/VIC11627/2020\|EPI_ISL_563437\|202 | ···················· | ···················· | ···················· | ···················· | ·····R·············· | ···················· | |
| K | 4102 | hCoV-19/Australia/VIC11628/2020\|EPI_ISL_563438\|202 | ···················· | ···················· | ···················· | ···················· | ···················· | ···················· | |
| K | 4103 | hCoV-19/Australia/VIC11634/2020\|EPI_ISL_563439\|202 | ···················· | ···················· | ···················· | ···················· | ···················· | ···················· | |
| K | 4104 | hCoV-19/Australia/VIC11636/2020\|EPI_ISL_563440\|202 | ···················· | ···················· | ···················· | ···················· | ···················· | ···················· | |
| K | 4105 | hCoV-19/Australia/VIC11638/2020\|EPI_ISL_563441\|202 | ···················· | ···················· | ···················· | ···················· | ···················· | ···················· | |
| K | 4106 | hCoV-19/Australia/VIC11639/2020\|EPI_ISL_563442\|202 | ···················· | ···················· | ···················· | ···················· | ···················· | ···················· | |
| K | 4107 | hCoV-19/Australia/VIC11640/2020\|EPI_ISL_563443\|202 | ···················· | ···················· | ···················· | ···················· | ···················· | ···················· | |
| K | 4110 | hCoV-19/Australia/VIC11649/2020\|EPI_ISL_563446\|202 | ···················· | ···················· | ···················· | ···················· | ···················· | ···················· | |
| K | 4111 | hCoV-19/Australia/VIC11651/2020\|EPI_ISL_563447\|202 | ···················· | ···················· | ···················· | ···················· | ···················· | ···················· | |
| K | 4112 | hCoV-19/Australia/VIC11654/2020\|EPI_ISL_563448\|202 | ···················· | ···················· | ···················· | ···················· | ···················· | ···················· | |
| K | 4113 | hCoV-19/Australia/VIC11658/2020\|EPI_ISL_563449\|202 | ···················· | ···················· | ···················· | ···················· | ···················· | ···················· | |
| K | 4115 | hCoV-19/Australia/VIC11665/2020\|EPI_ISL_563451\|202 | ···················· | ···················· | ···················· | ···················· | ···················· | ···················· | |
| K | 4116 | hCoV-19/Australia/VIC11667/2020\|EPI_ISL_563452\|202 | ···················· | ···················· | ···················· | ···················· | ···················· | ···················· | |
| K | 4117 | hCoV-19/Australia/VIC11670/2020\|EPI_ISL_563453\|202 | ···················· | ···················· | ···················· | ···················· | ···················· | ···················· | |
| K | 4118 | hCoV-19/Australia/VIC11672/2020\|EPI_ISL_563454\|202 | ···················· | ···················· | ···················· | ···················· | ···················· | ···················· | |
| K | 4119 | hCoV-19/Australia/VIC11677/2020\|EPI_ISL_563455\|202 | ···················· | ···G················ | ···················· | ···················· | ···················· | ·N·················· | |
| K | 4121 | hCoV-19/Australia/VIC11680/2020\|EPI_ISL_563457\|202 | ···················· | ···················· | ···················· | ···················· | ···················· | ···················· | |
| K | 4122 | hCoV-19/Australia/VIC11681/2020\|EPI_ISL_563458\|202 | ···················· | ···················· | ···················· | ···················· | ···················· | ···················· | |
| K | 4123 | hCoV-19/Australia/VIC11682/2020\|EPI_ISL_563459\|202 | ···················· | ···················· | ···················· | ···················· | ···················· | ···················· | |
| K | 4124 | hCoV-19/Australia/VIC11693/2020\|EPI_ISL_563462\|202 | ···················· | ···················· | ···················· | ···················· | ···················· | ···················· | |
| K | 4126 | hCoV-19/Australia/VIC11708/2020\|EPI_ISL_563467\|202 | ···················· | ···················· | ···················· | ···················· | ···················· | NNN················· | 1 |
| K | 4127 | hCoV-19/Australia/VIC11710/2020\|EPI_ISL_563468\|202 | ···················· | ···················· | ···················· | ···················· | ···················· | ···················· | |
| K | 4128 | hCoV-19/Australia/VIC11711/2020\|EPI_ISL_563469\|202 | ···················· | ···················· | ···················· | ···················· | ···················· | ···················· | |
| K | 4129 | hCoV-19/Australia/VIC11721/2020\|EPI_ISL_563470\|202 | ···················· | ···················· | ···················· | ···················· | ···················· | ···················· | |
| K | 4130 | hCoV-19/Australia/VIC11722/2020\|EPI_ISL_563471\|202 | ···················· | ···················· | ···················· | ···················· | ···················· | ···················· | |
| K | 4131 | hCoV-19/Australia/VIC11726/2020\|EPI_ISL_563472\|202 | ···················· | ···················· | ···················· | ···················· | ···················· | ···················· | |
| K | 4132 | hCoV-19/Australia/VIC11728/2020\|EPI_ISL_563473\|202 | ···················· | ···················· | ···················· | ···················· | ···················· | ···················· | |
| K | 4133 | hCoV-19/Australia/VIC11733/2020\|EPI_ISL_563474\|202 | ···················· | ···················· | ···················· | ···················· | ···················· | ···················· | |
| K | 4134 | hCoV-19/Australia/VIC11734/2020\|EPI_ISL_563475\|202 | ···················· | ···················· | ···················· | ···················· | ···················· | ···················· | |
| K | 4135 | hCoV-19/Australia/VIC11735/2020\|EPI_ISL_563476\|202 | ···················· | ···················· | ···················· | ···················· | ···················· | ···················· | |
| K | 4136 | hCoV-19/Australia/VIC7948/2020\|EPI_ISL_564764\|2020 | ···················· | ···················· | ···················· | ···················· | ···················· | ···················· | |
| K | 4137 | hCoV-19/Australia/VIC7951/2020\|EPI_ISL_564765\|2020 | ···················· | ···················· | ···················· | ···················· | ···················· | ···················· | |
| K | 4138 | hCoV-19/Australia/VIC7955/2020\|EPI_ISL_564766\|2020 | ···················· | ···················· | ···················· | ···················· | ···················· | ···················· | |
| K | 4139 | hCoV-19/Australia/VIC7956/2020\|EPI_ISL_564767\|2020 | ···················· | ···················· | ···················· | ···················· | ···················· | ···················· | |
| K | 4140 | hCoV-19/Australia/VIC7957/2020\|EPI_ISL_564768\|2020 | ···················· | ···················· | ···················· | ···················· | ···················· | ···················· | |
| K | 4141 | hCoV-19/Australia/VIC7958/2020\|EPI_ISL_564769\|2020 | ···················· | ···················· | ···················· | ···················· | ···················· | ···················· | |
| K | 4142 | hCoV-19/Australia/VIC7959/2020\|EPI_ISL_564770\|2020 | ···················· | ···················· | ···················· | ···················· | ···················· | ···················· | |
| K | 4143 | hCoV-19/Australia/VIC7960/2020\|EPI_ISL_564771\|2020 | ···················· | ···················· | ···················· | ···················· | ···················· | ···················· | |
| K | 4144 | hCoV-19/Australia/VIC7961/2020\|EPI_ISL_564772\|2020 | ···················· | ···················· | ···················· | ···················· | ···················· | ···················· | |
| K | 4145 | hCoV-19/Australia/VIC7962/2020\|EPI_ISL_564773\|2020 | ···················· | ···················· | ···················· | ···················· | ···················· | ···················· | |
| K | 4146 | hCoV-19/Australia/VIC7963/2020\|EPI_ISL_564774\|2020 | ···················· | ···················· | ···················· | ···················· | ···················· | ···················· | |
| K | 4147 | hCoV-19/Australia/VIC7964/2020\|EPI_ISL_564775\|2020 | ···················· | ···················· | ···················· | ···················· | ···················· | ···················· | |
| K | 4148 | hCoV-19/Australia/VIC7966/2020\|EPI_ISL_564776\|2020 | ···················· | ···················· | ···················· | ···················· | ···················· | ···················· | |
| K | 4149 | hCoV-19/Australia/VIC7968/2020\|EPI_ISL_564777\|2020 | ···················· | ···················· | ···················· | ···················· | ···················· | ···················· | |
| K | 4150 | hCoV-19/Australia/VIC7969/2020\|EPI_ISL_564778\|2020 | ···················· | ···················· | ···················· | ···················· | ···················· | ···················· | |
| K | 4151 | hCoV-19/Australia/VIC7970/2020\|EPI_ISL_564779\|2020 | ···················· | ···················· | ···················· | ···················· | ···················· | ···················· | |
| K | 4152 | hCoV-19/Australia/VIC7971/2020\|EPI_ISL_564780\|2020 | ···················· | ···················· | ···················· | ···················· | ···················· | ···················· | |
| K | 4153 | hCoV-19/Australia/VIC7973/2020\|EPI_ISL_564781\|2020 | ···················· | ···················· | ···················· | ···················· | ···················· | ···················· | |
| K | 4154 | hCoV-19/Australia/VIC7974/2020\|EPI_ISL_564782\|2020 | ···················· | ···················· | ···················· | ···················· | ···················· | ···················· | |
| K | 4155 | hCoV-19/Australia/VIC7976/2020\|EPI_ISL_564783\|2020 | ···················· | ···················· | ···················· | ···················· | ···················· | ···················· | |
| K | 4156 | hCoV-19/Australia/VIC7977/2020\|EPI_ISL_564784\|2020 | ···················· | ···················· | ···················· | ···················· | ···················· | ···················· | |
| K | 4157 | hCoV-19/Australia/VIC7978/2020\|EPI_ISL_564785\|2020 | ···················· | ···················· | ···················· | ···················· | ···················· | ···················· | |
| K | 4158 | hCoV-19/Australia/VIC7979/2020\|EPI_ISL_564786\|2020 | ···················· | ···················· | ···················· | ···················· | ···················· | ···················· | |
| K | 4159 | hCoV-19/Australia/VIC7980/2020\|EPI_ISL_564787\|2020 | ···················· | ···················· | ···················· | ···················· | ···················· | ···················· | |
| K | 4160 | hCoV-19/Australia/VIC7982/2020\|EPI_ISL_564788\|2020 | ···················· | ···················· | ···················· | ···················· | ···················· | ···················· | |
| K | 4161 | hCoV-19/Australia/VIC7984/2020\|EPI_ISL_564789\|2020 | ···················· | ···················· | ···················· | ···················· | ···················· | ···················· | |
| K | 4162 | hCoV-19/Australia/VIC7988/2020\|EPI_ISL_564793\|2020 | ···················· | ···················· | ···················· | ···················· | ···················· | ···················· | |
| K | 4163 | hCoV-19/Australia/VIC7997/2020\|EPI_ISL_564797\|2020 | ···················· | ···················· | ···················· | ···················· | ···················· | ···················· | |
| K | 4164 | hCoV-19/Australia/VIC7998/2020\|EPI_ISL_564798\|2020 | ···················· | ···················· | ···················· | ···················· | ···················· | ···················· | |
| K | 4165 | hCoV-19/Australia/VIC8006/2020\|EPI_ISL_564799\|2020 | ···················· | ···················· | ···················· | ···················· | ···················· | ···················· | |
| K | 4166 | hCoV-19/England/ALDP-9BEB52/2020\|EPI_ISL_568200\|20 | ···················· | ···················· | ···················· | ···················· | ···················· | ···················· | |
| K | 4167 | hCoV-19/England/ALDP-9BE211/2020\|EPI_ISL_568201\|20 | ···················· | ·A·················· | ···················· | ···················· | ···················· | ···A················ | |
| K | 4168 | hCoV-19/England/ALDP-9BE3A5/2020\|EPI_ISL_568202\|20 | ···················· | ···················· | ···················· | ···················· | ···················· | ···················· | |
| K | 4169 | hCoV-19/England/ALDP-9BDB9F/2020\|EPI_ISL_568203\|20 | ···················· | ···················· | ···················· | ···················· | ···················· | ···················· | |
| K | 4170 | hCoV-19/England/ALDP-9BE0BD/2020\|EPI_ISL_568205\|20 | ···················· | ···················· | ···················· | ···················· | ···················· | ···················· | |

| | | | | | | | | |
|---|---|---|---|---|---|---|---|---|
| K | 4171 | hCoV-19/England/ALDP-9BE6AC/2020\|EPI_ISL_568206\|20 | ···················· | ···················· | ····················· | ···················· | ···················· | ···················· |
| K | 4172 | hCoV-19/Scotland/QEUH-9B9BA2/2020\|EPI_ISL_568229\|2 | ···················· | ···················· | ····················· | ···················· | ···················· | ···················· |
| K | 4173 | hCoV-19/Scotland/QEUH-9B896D/2020\|EPI_ISL_568230\|2 | ···················· | ···················· | ····················· | ···················· | ···················· | ···················· |
| K | 4174 | hCoV-19/Scotland/QEUH-9B86FD/2020\|EPI_ISL_568231\|2 | ···················· | ···················· | ····················· | ···················· | ···················· | ···················· |
| K | 4175 | hCoV-19/Scotland/QEUH-9B9B75/2020\|EPI_ISL_568232\|2 | ···················· | ···················· | ····················· | ···················· | ···················· | ···················· |
| K | 4176 | hCoV-19/Scotland/QEUH-9B8F3E/2020\|EPI_ISL_568233\|2 | ···················· | ···················· | ····················· | ···················· | ···················· | ···················· |
| K | 4177 | hCoV-19/Scotland/QEUH-9B9B84/2020\|EPI_ISL_568234\|2 | ···················· | ···················· | ····················· | ···················· | ···················· | ···················· |
| K | 4178 | hCoV-19/Scotland/QEUH-9B82BD/2020\|EPI_ISL_568235\|2 | ···················· | ···················· | ····················· | ···················· | ···················· | ···················· |
| K | 4179 | hCoV-19/Scotland/QEUH-9B8D34/2020\|EPI_ISL_568236\|2 | ···················· | ···················· | ····················· | ···················· | ···················· | ···················· |
| K | 4180 | hCoV-19/Scotland/QEUH-9B82AE/2020\|EPI_ISL_568237\|2 | ···················· | ···················· | ····················· | ···················· | ···················· | ···················· |
| K | 4181 | hCoV-19/Scotland/QEUH-9B87CD/2020\|EPI_ISL_568238\|2 | ···················· | ···················· | ····················· | ···················· | ···················· | ···················· |
| K | 4182 | hCoV-19/Scotland/QEUH-9B9D8E/2020\|EPI_ISL_568239\|2 | ···················· | ···················· | ····················· | ···················· | ···················· | ···················· |
| K | 4183 | hCoV-19/Scotland/QEUH-9B845D/2020\|EPI_ISL_568240\|2 | ···················· | ···················· | ····················· | ···················· | ···················· | ···················· |
| K | 4184 | hCoV-19/Scotland/QEUH-9B84C6/2020\|EPI_ISL_568250\|2 | ···················· | ···················· | ····················· | ···················· | ···················· | ···················· |
| K | 4185 | hCoV-19/Scotland/QEUH-9B846C/2020\|EPI_ISL_568251\|2 | ···················· | ···················· | ····················· | ···················· | ···················· | ···················· |
| K | 4186 | hCoV-19/Scotland/QEUH-9B9A69/2020\|EPI_ISL_568252\|2 | ···················· | ···················· | ····················· | ···················· | ···················· | ···················· |
| K | 4187 | hCoV-19/Scotland/QEUH-9B9A5A/2020\|EPI_ISL_568253\|2 | ···················· | ···················· | ····················· | ···················· | ···················· | ···················· |
| K | 4188 | hCoV-19/Scotland/QEUH-9B88AC/2020\|EPI_ISL_568254\|2 | ···················· | ···················· | ····················· | ···················· | ···················· | ···················· |
| K | 4189 | hCoV-19/Scotland/QEUH-9B8253/2020\|EPI_ISL_568255\|2 | ···················· | ···················· | ····················· | ···················· | ···················· | ···················· |
| K | 4190 | hCoV-19/Scotland/QEUH-9B9C63/2020\|EPI_ISL_568256\|2 | ···················· | ···················· | ····················· | ···················· | ···················· | ···················· |
| K | 4191 | hCoV-19/Scotland/QEUH-9B85E1/2020\|EPI_ISL_568257\|2 | ···················· | ···················· | ····················· | ···················· | ···················· | ···················· |
| K | 4192 | hCoV-19/Scotland/QEUH-9B9DAC/2020\|EPI_ISL_568258\|2 | ···················· | ···················· | ····················· | ···················· | ···················· | ···················· |
| K | 4193 | hCoV-19/Scotland/QEUH-9B9AA5/2020\|EPI_ISL_568259\|2 | ···················· | ···················· | ····················· | ···················· | ···················· | ···················· |
| K | 4194 | hCoV-19/Scotland/QEUH-9B8ED7/2020\|EPI_ISL_568260\|2 | ···················· | ···················· | ····················· | ···················· | ···················· | ···················· |
| K | 4195 | hCoV-19/Scotland/QEUH-9B8314/2020\|EPI_ISL_568261\|2 | ···················· | ···················· | ····················· | ···················· | ···················· | ···················· |
| K | 4196 | hCoV-19/Scotland/QEUH-9B82DB/2020\|EPI_ISL_568262\|2 | ···················· | ···················· | ····················· | ···················· | ···················· | ···················· |
| K | 4197 | hCoV-19/Scotland/QEUH-9B8235/2020\|EPI_ISL_568263\|2 | ···················· | ···················· | ····················· | ···················· | ···················· | ···················· |
| K | 4198 | hCoV-19/Scotland/QEUH-9B9DCA/2020\|EPI_ISL_568264\|2 | ···················· | ···················· | ····················· | ···················· | ···················· | ···················· |
| K | 4199 | hCoV-19/Scotland/QEUH-9B8217/2020\|EPI_ISL_568265\|2 | ···················· | ···················· | ····················· | ···················· | ···················· | ···················· |
| K | 4200 | hCoV-19/Scotland/QEUH-9B8244/2020\|EPI_ISL_568266\|2 | ···················· | ···················· | ····················· | ···················· | ···················· | ···················· |
| K | 4201 | hCoV-19/Scotland/QEUH-9B9BB1/2020\|EPI_ISL_568267\|2 | ···················· | ···················· | ····················· | ···················· | ···················· | ···················· |
| K | 4202 | hCoV-19/Scotland/QEUH-9B8736/2020\|EPI_ISL_568268\|2 | ···················· | ···················· | ····················· | ···················· | ···················· | ···················· |
| K | 4203 | hCoV-19/Scotland/QEUH-9B86EE/2020\|EPI_ISL_568269\|2 | ···················· | ···················· | ····················· | ···················· | ···················· | ···················· |
| K | 4204 | hCoV-19/Scotland/QEUH-9B84B7/2020\|EPI_ISL_568270\|2 | ···················· | ······C············· | ····················· | ···················· | ···················· | ···················· |
| K | 4205 | hCoV-19/Scotland/QEUH-9B85D2/2020\|EPI_ISL_568271\|2 | ···················· | ···················· | ····················· | ···················· | ···················· | ···················· |
| K | 4206 | hCoV-19/Scotland/QEUH-9B8305/2020\|EPI_ISL_568272\|2 | ···················· | ···················· | ····················· | ···················· | ···················· | ···················· |
| K | 4207 | hCoV-19/Scotland/QEUH-9B84F3/2020\|EPI_ISL_568273\|2 | ···················· | ···················· | ····················· | ···················· | ···················· | ···················· |
| K | 4208 | hCoV-19/Scotland/QEUH-9B9AB4/2020\|EPI_ISL_568274\|2 | ···················· | ···················· | ····················· | ···················· | ···················· | ···················· |
| K | 4209 | hCoV-19/Scotland/QEUH-9B84D5/2020\|EPI_ISL_568275\|2 | ···················· | ···················· | ····················· | ···················· | ···················· | ···················· |
| K | 4210 | hCoV-19/Scotland/QEUH-9B9C72/2020\|EPI_ISL_568276\|2 | ···················· | ···················· | ····················· | ···················· | ···················· | ···················· |
| K | 4211 | hCoV-19/Scotland/QEUH-9B8718/2020\|EPI_ISL_568277\|2 | ···················· | ···················· | ····················· | ···················· | ···················· | ···················· |
| K | 4212 | hCoV-19/Scotland/QEUH-9B8262/2020\|EPI_ISL_568278\|2 | ···················· | ···················· | ····················· | ···················· | ···················· | ···················· |
| K | 4213 | hCoV-19/Scotland/QEUH-9B8208/2020\|EPI_ISL_568279\|2 | ···················· | ···················· | ····················· | ···················· | ···················· | ···················· |
| K | 4214 | hCoV-19/Scotland/QEUH-9B9A87/2020\|EPI_ISL_568281\|2 | ···················· | ·A·················· | ····················· | ···················· | ···················· | ···················· |
| K | 4215 | hCoV-19/Scotland/QEUH-9B9E5E/2020\|EPI_ISL_568282\|2 | ···················· | ···················· | ····················· | ···················· | ···················· | ···················· |
| K | 4216 | hCoV-19/Scotland/QEUH-9B8D25/2020\|EPI_ISL_568283\|2 | ···················· | ···················· | ····················· | ···················· | ···················· | ···················· |
| K | 4217 | hCoV-19/Scotland/QEUH-9B89C7/2020\|EPI_ISL_568284\|2 | ···················· | ···················· | ····················· | ···················· | ···················· | ···················· |
| K | 4218 | hCoV-19/Scotland/QEUH-9B8B0D/2020\|EPI_ISL_568285\|2 | ···················· | ···················· | ····················· | ···················· | ···················· | ···················· |
| K | 4219 | hCoV-19/Scotland/QEUH-9B860C/2020\|EPI_ISL_568286\|2 | ···················· | ···················· | ····················· | ···················· | ···················· | ···················· |
| K | 4220 | hCoV-19/Scotland/QEUH-9B8271/2020\|EPI_ISL_568287\|2 | ···················· | ···················· | ····················· | ···················· | ···················· | ···················· |
| K | 4221 | hCoV-19/Scotland/QEUH-9B9D24/2020\|EPI_ISL_568288\|2 | ···················· | ···················· | ····················· | ···················· | ···················· | ···················· |
| K | 4222 | hCoV-19/England/MILK-9A6BB7/2020\|EPI_ISL_568289\|20 | ···················· | ···················· | ····················· | ···················· | ···················· | ···················· |
| K | 4223 | hCoV-19/France/PAC-IHU-0428/2020\|EPI_ISL_569500\|20 | ···················· | ···················· | ····················· | ···················· | ···················· | ···················· |
| K | 4224 | hCoV-19/France/PAC-IHU-0429/2020\|EPI_ISL_569501\|20 | ···················· | ···················· | ····················· | ···················· | ···················· | ···················· |
| K | 4225 | hCoV-19/France/PAC-IHU-0430/2020\|EPI_ISL_569502\|20 | ···················· | ···················· | ····················· | ···················· | ···················· | ···················· |
| K | 4226 | hCoV-19/France/PAC-IHU-0431/2020\|EPI_ISL_569503\|20 | ···················· | ···················· | ····················· | ···················· | ···················· | ···················· |
| K | 4227 | hCoV-19/France/PAC-IHU-0432/2020\|EPI_ISL_569504\|20 | ···················· | ···················· | ····················· | ···················· | ···················· | ···················· |
| K | 4228 | hCoV-19/France/PAC-IHU-0433/2020\|EPI_ISL_569505\|20 | ···················· | ···················· | ····················· | ···················· | ···················· | ···················· |
| K | 4229 | hCoV-19/France/PAC-IHU-0434/2020\|EPI_ISL_569506\|20 | ···················· | ···················· | ····················· | ···················· | ···················· | ···················· |
| K | 4230 | hCoV-19/France/PAC-IHU-0435/2020\|EPI_ISL_569507\|20 | ···················· | ···················· | ····················· | ···················· | ···················· | ···················· |
| K | 4231 | hCoV-19/France/PAC-IHU-0438/2020\|EPI_ISL_569508\|20 | ···················· | ···················· | ····················· | ···················· | ···················· | ···················· |
| K | 4232 | hCoV-19/France/PAC-IHU-0439/2020\|EPI_ISL_569509\|20 | ···················· | ···················· | ····················· | ···················· | ···················· | ···················· |
| K | 4233 | hCoV-19/France/PAC-IHU-0440/2020\|EPI_ISL_569510\|20 | ···················· | ···················· | ····················· | ···················· | ···················· | ···················· |
| K | 4234 | hCoV-19/France/PAC-IHU-0441/2020\|EPI_ISL_569511\|20 | ···················· | ···················· | ····················· | ···················· | ···················· | ···················· |
| K | 4235 | hCoV-19/France/PAC-IHU-0444/2020\|EPI_ISL_569512\|20 | ···················· | ···················· | ····················· | ···················· | ···················· | ···················· |
| K | 4236 | hCoV-19/France/PAC-IHU-0445/2020\|EPI_ISL_569513\|20 | ···················· | ···················· | ····················· | ···················· | ···················· | ···················· |
| K | 4237 | hCoV-19/France/PAC-IHU-0446/2020\|EPI_ISL_569514\|20 | ···················· | ···················· | ····················· | ···················· | ···················· | ···················· |
| K | 4238 | hCoV-19/France/PAC-IHU-0447/2020\|EPI_ISL_569515\|20 | ···················· | ···················· | ····················· | ···················· | ···················· | ···················· |
| K | 4239 | hCoV-19/France/PAC-IHU-0449/2020\|EPI_ISL_569516\|20 | ···················· | ···················· | ····················· | ···················· | ···················· | ···················· |
| K | 4240 | hCoV-19/France/PAC-IHU-0450/2020\|EPI_ISL_569517\|20 | ···················· | ···················· | ····················· | ···················· | ···················· | ···················· |
| K | 4241 | hCoV-19/France/PAC-IHU-0451/2020\|EPI_ISL_569518\|20 | ···················· | ···················· | ····················· | ···················· | ···················· | ···················· |
| K | 4242 | hCoV-19/France/PAC-IHU-0452/2020\|EPI_ISL_569519\|20 | ···················· | ···················· | ····················· | ···················· | ···················· | ···················· |
| K | 4243 | hCoV-19/France/PAC-IHU-0453/2020\|EPI_ISL_569520\|20 | ···················· | ···················· | ····················· | ···················· | ···················· | ···················· |
| K | 4244 | hCoV-19/France/PAC-IHU-0455/2020\|EPI_ISL_569521\|20 | ···················· | ···················· | ····················· | ···················· | ···················· | ···················· |
| K | 4245 | hCoV-19/France/PAC-IHU-0456/2020\|EPI_ISL_569522\|20 | ···················· | ···················· | ····················· | ···················· | ···················· | ···················· |
| K | 4246 | hCoV-19/France/PAC-IHU-0457/2020\|EPI_ISL_569523\|20 | ···················· | ···················· | ····················· | ···················· | ···················· | ···················· |
| K | 4247 | hCoV-19/France/PAC-IHU-0459/2020\|EPI_ISL_569524\|20 | ···················· | ···················· | ····················· | ···················· | ···················· | ···················· |
| K | 4248 | hCoV-19/France/PAC-IHU-0460/2020\|EPI_ISL_569525\|20 | ···················· | ···················· | ····················· | ···················· | ···················· | ···················· |
| K | 4249 | hCoV-19/France/PAC-IHU-0461/2020\|EPI_ISL_569526\|20 | ···················· | ···················· | ····················· | ···················· | ···················· | ···················· |
| K | 4250 | hCoV-19/France/PAC-IHU-0462/2020\|EPI_ISL_569527\|20 | ···················· | ···················· | ····················· | ···················· | ···················· | ···················· |

| | | | | | | | | |
|---|---|---|---|---|---|---|---|---|
| K | 4251 | hCoV-19/France/PAC-IHU-0464/2020\|EPI_ISL_569528\|20 | ···················· | ···················· | ···················· | ···················· | ···················· | ···················· |
| K | 4252 | hCoV-19/France/PAC-IHU-0465/2020\|EPI_ISL_569529\|20 | ···················· | ···················· | ···················· | ···················· | ···················· | ···················· |
| K | 4253 | hCoV-19/France/PAC-IHU-0466/2020\|EPI_ISL_569530\|20 | ···················· | ···················· | ···················· | ···················· | ···················· | ···················· |
| K | 4254 | hCoV-19/France/PAC-IHU-0467/2020\|EPI_ISL_569531\|20 | ···················· | ···················· | ···················· | ···················· | ···················· | ···················· |
| K | 4255 | hCoV-19/France/PAC-IHU-0468/2020\|EPI_ISL_569532\|20 | ···················· | ···················· | ···················· | ···················· | ···················· | ···················· |
| K | 4256 | hCoV-19/France/PAC-IHU-0469/2020\|EPI_ISL_569533\|20 | ···················· | ···················· | ···················· | ···················· | ···················· | ···················· |
| K | 4257 | hCoV-19/France/PAC-IHU-0470/2020\|EPI_ISL_569534\|20 | ···················· | ···················· | ···················· | ···················· | ···················· | ···················· |
| K | 4258 | hCoV-19/France/PAC-IHU-0471/2020\|EPI_ISL_569535\|20 | ···················· | ···················· | ···················· | ···················· | ···················· | ···················· |
| K | 4259 | hCoV-19/France/PAC-IHU-0472/2020\|EPI_ISL_569536\|20 | ···················· | ···················· | ···················· | ···················· | ···················· | ···················· |
| K | 4260 | hCoV-19/France/PAC-IHU-0473/2020\|EPI_ISL_569537\|20 | ···················· | ···················· | ···················· | ···················· | ···················· | ···················· |
| K | 4261 | hCoV-19/France/PAC-IHU-0506/2020\|EPI_ISL_569560\|20 | ···················· | ···················· | ···················· | ···················· | ···················· | ···················· |
| K | 4262 | hCoV-19/France/PAC-IHU-0507/2020\|EPI_ISL_569561\|20 | ···················· | ···················· | ···················· | ···················· | ···················· | ···················· |
| K | 4263 | hCoV-19/France/PAC-IHU-0508/2020\|EPI_ISL_569562\|20 | ···················· | ···················· | ···················· | ···················· | ···················· | ···················· |
| K | 4264 | hCoV-19/France/PAC-IHU-0510/2020\|EPI_ISL_569563\|20 | ···················· | ···················· | ···················· | ···················· | ···················· | ···················· |
| K | 4265 | hCoV-19/France/PAC-IHU-0511/2020\|EPI_ISL_569564\|20 | ···················· | ···················· | ···················· | ···················· | ···················· | ···················· |
| K | 4266 | hCoV-19/France/PAC-IHU-0512/2020\|EPI_ISL_569565\|20 | ···················· | ···················· | ···················· | ···················· | ···················· | ···················· |
| K | 4267 | hCoV-19/France/PAC-IHU-0516/2020\|EPI_ISL_569566\|20 | ···················· | ···················· | ···················· | ···················· | ···················· | ···················· |
| K | 4268 | hCoV-19/France/PAC-IHU-0518/2020\|EPI_ISL_569567\|20 | ···················· | ···················· | ···················· | ···················· | ···················· | ···················· |
| K | 4269 | hCoV-19/France/PAC-IHU-0520/2020\|EPI_ISL_569568\|20 | ···················· | ···················· | ···················· | ···················· | ···················· | ···················· |
| K | 4270 | hCoV-19/France/PAC-IHU-0521/2020\|EPI_ISL_569569\|20 | ···················· | ···················· | ···················· | ···················· | ···················· | ···················· |
| K | 4271 | hCoV-19/France/PAC-IHU-0525/2020\|EPI_ISL_569571\|20 | ···················· | ···················· | ···················· | ···················· | ···················· | ···················· |
| K | 4272 | hCoV-19/France/PAC-IHU-0540/2020\|EPI_ISL_569582\|20 | ···················· | ···················· | ···················· | ···················· | ···················· | ···················· |
| K | 4273 | hCoV-19/France/PAC-IHU-0542/2020\|EPI_ISL_569583\|20 | ···················· | ···················· | ···················· | ···················· | ···················· | ···················· |
| K | 4274 | hCoV-19/France/PAC-IHU-0545/2020\|EPI_ISL_569584\|20 | ···················· | ···················· | ···················· | ···················· | ···················· | ···················· |
| K | 4275 | hCoV-19/France/PAC-IHU-0547/2020\|EPI_ISL_569585\|20 | ···················· | ···················· | ···················· | ···················· | ···················· | ···················· |
| K | 4276 | hCoV-19/France/PAC-IHU-0548/2020\|EPI_ISL_569586\|20 | ···················· | ···················· | ···················· | ···················· | ···················· | ···················· |
| K | 4277 | hCoV-19/France/PAC-IHU-0549/2020\|EPI_ISL_569587\|20 | ···················· | ···················· | ···················· | ···················· | ···················· | ···················· |
| K | 4278 | hCoV-19/France/PAC-IHU-0550/2020\|EPI_ISL_569588\|20 | ···················· | ···················· | ···················· | ···················· | ···················· | ···················· |
| K | 4279 | hCoV-19/France/PAC-IHU-0551/2020\|EPI_ISL_569589\|20 | ···················· | ···················· | ···················· | ···················· | ···················· | ···················· |
| K | 4280 | hCoV-19/France/PAC-IHU-0553/2020\|EPI_ISL_569590\|20 | ···················· | ···················· | ···················· | ···················· | ···················· | ···················· |
| K | 4281 | hCoV-19/France/PAC-IHU-0555/2020\|EPI_ISL_569592\|20 | ···················· | ···················· | ···················· | ···················· | ···················· | ···················· |
| K | 4282 | hCoV-19/France/PAC-IHU-0558/2020\|EPI_ISL_569593\|20 | ···················· | ···················· | ···················· | ···················· | ···················· | ···················· |
| K | 4283 | hCoV-19/France/PAC-IHU-0559/2020\|EPI_ISL_569594\|20 | ···················· | ···················· | ···················· | ···················· | ···················· | ···················· |
| K | 4284 | hCoV-19/France/PAC-IHU-0560/2020\|EPI_ISL_569595\|20 | ···················· | ···················· | ···················· | ···················· | ···················· | ···················· |
| K | 4285 | hCoV-19/France/PAC-IHU-0573/2020\|EPI_ISL_569597\|20 | ···················· | ···················· | ···················· | ···················· | ···················· | ···················· |
| K | 4286 | hCoV-19/France/PAC-IHU-0575/2020\|EPI_ISL_569598\|20 | ···················· | ···················· | ···················· | ···················· | ···················· | ···················· |
| K | 4287 | hCoV-19/France/PAC-IHU-0576/2020\|EPI_ISL_569599\|20 | ···················· | ···················· | ···················· | ···················· | ···················· | ···················· |
| K | 4288 | hCoV-19/USA/WA-UW-12171/2020\|EPI_ISL_570500\|2020-0 | ···················· | ···················· | ···················· | ···················· | ···················· | ···················· |
| K | 4289 | hCoV-19/USA/WA-UW-12173/2020\|EPI_ISL_570501\|2020-0 | ···················· | ·A·················· | ···················· | ···················· | ···················· | ···················· |
| K | 4290 | hCoV-19/USA/WA-UW-12283/2020\|EPI_ISL_570502\|2020-0 | ···················· | ···················· | ···················· | ···················· | ···················· | ···················· |
| K | 4291 | hCoV-19/USA/WA-UW-12324/2020\|EPI_ISL_570503\|2020-0 | ···················· | ···················· | ···················· | ···················· | ···················· | ···················· |
| K | 4292 | hCoV-19/USA/WA-UW-12332/2020\|EPI_ISL_570504\|2020-0 | ···················· | ·A·················· | ···················· | ···················· | ···················· | ···················· |
| K | 4293 | hCoV-19/USA/WA-UW-12334/2020\|EPI_ISL_570506\|2020-0 | ···················· | ···················· | ···················· | ···················· | ···················· | ···················· |
| K | 4294 | hCoV-19/USA/WA-UW-12340/2020\|EPI_ISL_570507\|2020-0 | ···················· | ·A·················· | ···················· | ···················· | ···················· | ···················· |
| K | 4295 | hCoV-19/USA/WA-UW-12348/2020\|EPI_ISL_570508\|2020-0 | ···················· | ···················· | ···················· | ···················· | ···················· | ···················· |
| K | 4296 | hCoV-19/USA/WA-UW-12352/2020\|EPI_ISL_570509\|2020-0 | ···················· | ···················· | ···················· | ···················· | ···················· | ···················· |
| K | 4297 | hCoV-19/USA/WA-UW-12355/2020\|EPI_ISL_570510\|2020-0 | ···················· | ···················· | ···················· | ···················· | ···················· | ···················· |
| K | 4298 | hCoV-19/USA/WA-UW-12356/2020\|EPI_ISL_570511\|2020-0 | ···················· | ···················· | ···················· | ···················· | ······T············· | ···················· |
| K | 4299 | hCoV-19/USA/WA-UW-12359/2020\|EPI_ISL_570512\|2020-0 | ···················· | ···················· | ···················· | ···················· | ···················· | ···················· |
| K | 4300 | hCoV-19/USA/WA-UW-12360/2020\|EPI_ISL_570513\|2020-0 | ···················· | ···················· | ···················· | ···················· | ···················· | ···················· |
| K | 4301 | hCoV-19/USA/WA-UW-12361/2020\|EPI_ISL_570514\|2020-0 | ···················· | ···················· | ···················· | ···················· | ···················· | ···················· |
| K | 4302 | hCoV-19/USA/WA-UW-12362/2020\|EPI_ISL_570515\|2020-0 | ···················· | ···················· | ···················· | ···················· | ···················· | ···················· |
| K | 4303 | hCoV-19/USA/WA-UW-12364/2020\|EPI_ISL_570516\|2020-0 | ···················· | ···················· | ···················· | ···················· | ···················· | ···················· |
| K | 4304 | hCoV-19/USA/WA-UW-12367/2020\|EPI_ISL_570517\|2020-0 | ···················· | ···················· | ···················· | ···················· | ···················· | ···················· |
| K | 4305 | hCoV-19/USA/WA-UW-12368/2020\|EPI_ISL_570518\|2020-0 | ···················· | ···················· | ···················· | ···················· | ···················· | ···················· |
| K | 4306 | hCoV-19/USA/WA-UW-12371/2020\|EPI_ISL_570519\|2020-0 | ···················· | ···················· | ···················· | ···················· | ···················· | ···················· |
| K | 4307 | hCoV-19/USA/WA-UW-12516/2020\|EPI_ISL_570540\|2020-0 | ···················· | ···················· | ···················· | ···················· | ···················· | ···················· |
| K | 4308 | hCoV-19/USA/WA-UW-12521/2020\|EPI_ISL_570541\|2020-0 | ···················· | ···················· | ···················· | ···················· | ···················· | ···················· |
| K | 4309 | hCoV-19/USA/WA-UW-12531/2020\|EPI_ISL_570542\|2020-0 | ···················· | ···················· | ···················· | ···················· | ···················· | ···················· |
| K | 4310 | hCoV-19/USA/WA-UW-12539/2020\|EPI_ISL_570543\|2020-0 | ···················· | ·A·················· | ···················· | ···················· | ···················· | ···················· |
| K | 4311 | hCoV-19/USA/WA-UW-12543/2020\|EPI_ISL_570545\|2020-0 | ···················· | ···················· | ···················· | ···················· | ···················· | ···················· |
| K | 4312 | hCoV-19/USA/WA-UW-12548/2020\|EPI_ISL_570546\|2020-0 | ···················· | ···················· | ···················· | ···················· | ···················· | ···················· |
| K | 4313 | hCoV-19/USA/WA-UW-1255/2020\|EPI_ISL_570547\|2020-03 | ···················· | ···················· | ···················· | ···················· | ···················· | ···················· |
| K | 4314 | hCoV-19/USA/WA-UW-22230/2020\|EPI_ISL_570561\|2020-0 | ···················· | ···················· | ···················· | ···················· | ···················· | ···················· |
| K | 4315 | hCoV-19/USA/WA-UW-22057/2020\|EPI_ISL_570562\|2020-0 | ···················· | ···················· | ···················· | ···················· | ···················· | ···················· |
| K | 4316 | hCoV-19/USA/WA-UW-22060/2020\|EPI_ISL_570563\|2020-0 | ···················· | ·A·················· | ···················· | ···················· | ···················· | ···················· |
| K | 4317 | hCoV-19/USA/WA-UW-22103/2020\|EPI_ISL_570564\|2020-0 | ···················· | ·A·················· | ···················· | ···················· | ···················· | ···················· |
| K | 4318 | hCoV-19/USA/WA-UW-22137/2020\|EPI_ISL_570565\|2020-0 | ···················· | ·A·················· | ···················· | ···················· | ···················· | ···················· |
| K | 4319 | hCoV-19/USA/WA-UW-22155/2020\|EPI_ISL_570566\|2020-0 | ···················· | ···················· | ···················· | ···················· | ···················· | ···················· |
| K | 4320 | hCoV-19/USA/WA-UW-22162/2020\|EPI_ISL_570567\|2020-0 | ···················· | ···················· | ···················· | ···················· | ···················· | ···················· |
| K | 4321 | hCoV-19/USA/WA-UW-22172/2020\|EPI_ISL_570568\|2020-0 | ···················· | ···················· | ···················· | ···················· | ···················· | ···················· |
| K | 4322 | hCoV-19/USA/WA-UW-23194/2020\|EPI_ISL_570569\|2020-0 | ···················· | ···················· | ···················· | ···················· | ···················· | ···················· |
| K | 4323 | hCoV-19/USA/WA-UW-22847/2020\|EPI_ISL_570570\|2020-0 | ···················· | ···················· | ···················· | ···················· | ···················· | ···················· |
| K | 4324 | hCoV-19/USA/WA-UW-22256/2020\|EPI_ISL_570571\|2020-0 | ···················· | ·A·················· | ···················· | ···················· | ···················· | ···················· |
| K | 4325 | hCoV-19/USA/WA-UW-22817/2020\|EPI_ISL_570572\|2020-0 | ···················· | ···················· | ···················· | ···················· | ···················· | ···················· |
| K | 4326 | hCoV-19/USA/WA-UW-24887/2020\|EPI_ISL_570573\|2020-0 | ···················· | ···················· | ···················· | ···················· | ···················· | ···················· |
| K | 4327 | hCoV-19/USA/WA-UW-27035/2020\|EPI_ISL_570574\|2020-0 | ···················· | ···················· | ···················· | ···················· | ···················· | ···················· |
| K | 4328 | hCoV-19/USA/WA-UW-24946/2020\|EPI_ISL_570575\|2020-0 | ···················· | ···················· | ···················· | ···················· | ···················· | ···················· |
| K | 4329 | hCoV-19/USA/WA-UW-22855/2020\|EPI_ISL_570576\|2020-0 | ···················· | ···················· | ···················· | ···················· | ···················· | ···················· |
| K | 4330 | hCoV-19/USA/WA-UW-26495/2020\|EPI_ISL_570577\|2020-0 | ···················· | ·A·················· | ···················· | ···················· | ···················· | ···················· |

| | | | | | | | | | |
|---|---|---|---|---|---|---|---|---|---|
| K | 4331 | hCoV-19/USA/WA-UW-22460/2020\|EPI_ISL_570578\|2020-0 | ···················· | ···················· | ····················· | ···················· | ···················· | ···················· | |
| K | 4332 | hCoV-19/USA/WA-UW-26865/2020\|EPI_ISL_570579\|2020-0 | ···················· | ···················· | ····················· | ···················· | ···················· | ···················· | |
| K | 4333 | hCoV-19/USA/WA-UW-24802/2020\|EPI_ISL_570580\|2020-0 | ···················· | ···················· | ····················· | ···················· | ···················· | ···················· | |
| K | 4334 | hCoV-19/USA/WA-UW-24809/2020\|EPI_ISL_570581\|2020-0 | ···················· | ···················· | ····················· | ···················· | ···················· | ···················· | |
| K | 4335 | hCoV-19/USA/WA-UW-22353/2020\|EPI_ISL_570582\|2020-0 | ···················· | ···················· | ····················· | ···················· | ···················· | ···················· | |
| K | 4336 | hCoV-19/USA/WA-UW-24920/2020\|EPI_ISL_570583\|2020-0 | ···················· | ···················· | ····················· | ···················· | ···················· | ···················· | |
| K | 4337 | hCoV-19/USA/OR-UW-24378/2020\|EPI_ISL_570584\|2020-0 | ···················· | ···················· | ····················· | ···················· | ···················· | ···················· | |
| K | 4338 | hCoV-19/USA/WA-UW-22056/2020\|EPI_ISL_570585\|2020-0 | ···················· | ···················· | ····················· | ···················· | ···················· | ···················· | |
| K | 4339 | hCoV-19/USA/WA-UW-22190/2020\|EPI_ISL_570586\|2020-0 | ···················· | ···················· | ····················· | ···················· | ···················· | ···················· | |
| K | 4340 | hCoV-19/USA/WA-UW-22205/2020\|EPI_ISL_570587\|2020-0 | ···················· | ···················· | ····················· | ···················· | ···················· | ···················· | |
| K | 4341 | hCoV-19/USA/WA-UW-22216/2020\|EPI_ISL_570588\|2020-0 | ···················· | ·A····A············· | ····················· | ···················· | ···················· | ···················· | |
| K | 4342 | hCoV-19/USA/WA-UW-22236/2020\|EPI_ISL_570589\|2020-0 | ···················· | ·A·················· | ····················· | ···················· | ···················· | ···················· | |
| K | 4343 | hCoV-19/USA/WA-UW-22271/2020\|EPI_ISL_570590\|2020-0 | ···················· | ·A·················· | ····················· | ···················· | ···················· | ···················· | |
| K | 4344 | hCoV-19/USA/WA-UW-22415/2020\|EPI_ISL_570591\|2020-0 | ···················· | ···················· | ····················· | ···················· | ···················· | ···················· | |
| K | 4345 | hCoV-19/USA/WA-UW-22521/2020\|EPI_ISL_570592\|2020-0 | ··················T· | ···················· | ····················· | ···················· | ···················· | ···················· | 1 |
| K | 4346 | hCoV-19/USA/WA-UW-22809/2020\|EPI_ISL_570593\|2020-0 | ···················· | ···················· | ····················· | ···················· | ···················· | ···················· | |
| K | 4347 | hCoV-19/USA/WA-UW-23335/2020\|EPI_ISL_570594\|2020-0 | ···················· | ·A·················· | ····················· | ···················· | ···················· | ···················· | |
| K | 4348 | hCoV-19/USA/WA-UW-23555/2020\|EPI_ISL_570595\|2020-0 | ···················· | ·A·················· | ····················· | ···················· | ···················· | ···················· | |
| K | 4349 | hCoV-19/USA/WA-UW-23564/2020\|EPI_ISL_570596\|2020-0 | ···················· | ·A·················· | ····················· | ···················· | ···················· | ···················· | |
| K | 4350 | hCoV-19/USA/WA-UW-23691/2020\|EPI_ISL_570597\|2020-0 | ···················· | ·A·················· | ····················· | ···················· | ···················· | ···················· | |
| K | 4351 | hCoV-19/USA/WA-UW-23752/2020\|EPI_ISL_570598\|2020-0 | ···················· | ·A·················· | ····················· | ···················· | ···················· | ···················· | |
| K | 4352 | hCoV-19/USA/WA-UW-23782/2020\|EPI_ISL_570599\|2020-0 | ···················· | ·A·················· | ····················· | ···················· | ···················· | ···················· | |
| K | 4353 | hCoV-19/Australia/VIC13571/2020\|EPI_ISL_562168\|202 | ···················· | ···················· | ····················· | ···················· | ···················· | ···················· | |
| K | 4354 | hCoV-19/Australia/VIC9121/2020\|EPI_ISL_562169\|2020 | ···················· | ···················· | ····················· | ···················· | ···················· | ···················· | |
| K | 4355 | hCoV-19/Australia/VIC8500/2020\|EPI_ISL_562170\|2020 | ···················· | ···················· | ····················· | ···················· | ···················· | ···················· | |
| K | 4356 | hCoV-19/Australia/VIC8617/2020\|EPI_ISL_562171\|2020 | ···················· | ···················· | ····················· | ···················· | ···················· | ···················· | |
| K | 4357 | hCoV-19/Australia/VIC10124/2020\|EPI_ISL_562172\|202 | ···················· | ···················· | ····················· | ···················· | ···················· | ···················· | |
| K | 4358 | hCoV-19/Australia/VIC10540/2020\|EPI_ISL_562173\|202 | ···················· | ···················· | ····················· | ···················· | ···················· | ···················· | |
| K | 4359 | hCoV-19/Australia/VIC9920/2020\|EPI_ISL_562174\|2020 | ···················· | ···················· | ····················· | ···················· | ···················· | ···················· | |
| K | 4360 | hCoV-19/Australia/VIC10127/2020\|EPI_ISL_562800\|202 | ···················· | ···················· | ····················· | ···················· | ···················· | ···················· | |
| K | 4361 | hCoV-19/Australia/VIC10130/2020\|EPI_ISL_562801\|202 | ···················· | ···················· | ····················· | ···················· | ···················· | ···················· | |
| K | 4363 | hCoV-19/Australia/VIC10135/2020\|EPI_ISL_562803\|202 | ···················· | ···················· | ··K·················· | ···················· | ···················· | ···················· | |
| K | 4364 | hCoV-19/Australia/VIC10136/2020\|EPI_ISL_562804\|202 | ···················· | ···················· | ··K·················· | ···················· | ···················· | ···················· | |
| K | 4365 | hCoV-19/Australia/VIC10137/2020\|EPI_ISL_562805\|202 | ···················· | ···················· | ····················· | ···················· | ···················· | ···················· | |
| K | 4366 | hCoV-19/Australia/VIC10142/2020\|EPI_ISL_562806\|202 | ···················· | ···················· | ··K·················· | ···················· | ···················· | ···················· | |
| K | 4367 | hCoV-19/Australia/VIC10144/2020\|EPI_ISL_562807\|202 | ···················· | ···················· | ··K·················· | ···················· | ···················· | ···················· | |
| K | 4368 | hCoV-19/Australia/VIC10147/2020\|EPI_ISL_562808\|202 | ···················· | ···················· | ····················· | ···················· | ···················· | ···················· | |
| K | 4369 | hCoV-19/Australia/VIC10149/2020\|EPI_ISL_562809\|202 | ···················· | ···················· | ··N·················· | ···················· | ···················· | ···················· | |
| K | 4370 | hCoV-19/Australia/VIC10155/2020\|EPI_ISL_562810\|202 | ···················· | ···················· | ····················· | ···················· | ···················· | ···················· | |
| K | 4371 | hCoV-19/Australia/VIC10157/2020\|EPI_ISL_562811\|202 | ···················· | ···················· | ····················· | ···················· | ···················· | ···················· | |
| K | 4372 | hCoV-19/Australia/VIC10165/2020\|EPI_ISL_562812\|202 | ···················· | ···················· | ····················· | ···················· | ···················· | ···················· | |
| K | 4373 | hCoV-19/Australia/VIC10167/2020\|EPI_ISL_562813\|202 | ···················· | ···················· | ····················· | ···················· | ···················· | ···················· | |
| K | 4374 | hCoV-19/Australia/VIC10170/2020\|EPI_ISL_562814\|202 | ···················· | ···················· | ····················· | ···················· | ···················· | ···················· | |
| K | 4375 | hCoV-19/Australia/VIC10174/2020\|EPI_ISL_562815\|202 | ···················· | ···················· | ····················· | ···················· | ···················· | ···················· | |
| K | 4376 | hCoV-19/Australia/VIC10177/2020\|EPI_ISL_562816\|202 | ···················· | ···················· | ····················· | ···················· | ···················· | ···················· | |
| K | 4377 | hCoV-19/Australia/VIC10182/2020\|EPI_ISL_562817\|202 | ···················· | ···················· | ····················· | ···················· | ···················· | ···················· | |
| K | 4378 | hCoV-19/Australia/VIC10187/2020\|EPI_ISL_562818\|202 | ···················· | ···················· | ····················· | ···················· | ···················· | ···················· | |
| K | 4381 | hCoV-19/Australia/VIC10192/2020\|EPI_ISL_562821\|202 | ···················· | ···················· | ····················· | ···················· | ···················· | ···················· | |
| K | 4382 | hCoV-19/Australia/VIC10200/2020\|EPI_ISL_562822\|202 | ···················· | ···················· | ····················· | ···················· | ···················· | ···················· | |
| K | 4383 | hCoV-19/Australia/VIC10202/2020\|EPI_ISL_562823\|202 | ···················· | ···················· | ····················· | ···················· | ···················· | ···················· | |
| K | 4384 | hCoV-19/Australia/VIC10212/2020\|EPI_ISL_562824\|202 | ···················· | ···················· | ····················· | ···················· | ···················· | ···················· | |
| K | 4386 | hCoV-19/Australia/VIC10216/2020\|EPI_ISL_562826\|202 | ···················· | ···················· | ····················· | ···················· | ···················· | ···················· | |
| K | 4388 | hCoV-19/Australia/VIC10219/2020\|EPI_ISL_562828\|202 | ···················· | ···················· | ····················· | ···················· | ···················· | ···················· | |
| K | 4389 | hCoV-19/Australia/VIC10221/2020\|EPI_ISL_562829\|202 | ···················· | ···················· | ····················· | ···················· | ···················· | ···················· | |
| K | 4390 | hCoV-19/Australia/VIC10223/2020\|EPI_ISL_562830\|202 | ···················· | ···················· | ····················· | ···················· | ···················· | ···················· | |
| K | 4391 | hCoV-19/Australia/VIC10227/2020\|EPI_ISL_562831\|202 | ···················· | ···················· | ····················· | ···················· | ···················· | ···················· | |
| K | 4392 | hCoV-19/Australia/VIC10229/2020\|EPI_ISL_562832\|202 | ···················· | ···················· | ····················· | ···················· | ···················· | ···················· | |
| K | 4394 | hCoV-19/Australia/VIC10234/2020\|EPI_ISL_562834\|202 | ···················· | ···················· | ····················· | ···················· | ···················· | ···················· | |
| K | 4395 | hCoV-19/Australia/VIC10235/2020\|EPI_ISL_562835\|202 | ···················· | ···················· | ····················· | ···················· | ···················· | ···················· | |
| K | 4396 | hCoV-19/Australia/VIC10238/2020\|EPI_ISL_562836\|202 | ···················· | ···················· | ····················· | ···················· | ···················· | ···················· | |
| K | 4397 | hCoV-19/Australia/VIC10242/2020\|EPI_ISL_562837\|202 | ···················· | ···················· | ····················· | ···················· | ···················· | ···················· | |
| K | 4398 | hCoV-19/Australia/VIC10244/2020\|EPI_ISL_562838\|202 | ···················· | ···················· | ····················· | ···················· | ···················· | ···················· | |
| K | 4399 | hCoV-19/Australia/VIC10245/2020\|EPI_ISL_562839\|202 | ···················· | ···················· | ····················· | ···················· | ···················· | ···················· | |
| K | 4400 | hCoV-19/Australia/VIC10247/2020\|EPI_ISL_562840\|202 | ···················· | ···················· | ··K·················· | ···················· | ···················· | ···················· | |
| K | 4401 | hCoV-19/Australia/VIC10249/2020\|EPI_ISL_562841\|202 | ···················· | ···················· | ··N·················· | ···················· | ···················· | ···················· | |
| K | 4402 | hCoV-19/Australia/VIC10250/2020\|EPI_ISL_562842\|202 | ···················· | ···················· | ····················· | ···················· | ···················· | ···················· | |
| K | 4403 | hCoV-19/Australia/VIC10254/2020\|EPI_ISL_562845\|202 | ···················· | ···················· | ··K·················· | ···················· | ···················· | ···················· | |
| K | 4404 | hCoV-19/Australia/VIC10263/2020\|EPI_ISL_562846\|202 | ···················· | ···················· | ····················· | ···················· | ···················· | ···················· | |
| K | 4405 | hCoV-19/Australia/VIC10265/2020\|EPI_ISL_562847\|202 | ···················· | ···················· | ····················· | ···················· | ···················· | ···················· | |
| K | 4406 | hCoV-19/Australia/VIC10279/2020\|EPI_ISL_562848\|202 | ···················· | ···················· | ····················· | ···················· | ···················· | ···················· | |
| K | 4407 | hCoV-19/Australia/VIC10280/2020\|EPI_ISL_562849\|202 | ···················· | ···················· | ····················· | ···················· | ···················· | ···················· | |
| K | 4408 | hCoV-19/Australia/VIC10281/2020\|EPI_ISL_562850\|202 | ···················· | ···················· | ····················· | ···················· | ···················· | ···················· | |
| K | 4409 | hCoV-19/Australia/VIC10283/2020\|EPI_ISL_562851\|202 | ···················· | ···················· | ····················· | ···················· | ···················· | ···················· | |
| K | 4411 | hCoV-19/Australia/VIC10285/2020\|EPI_ISL_562853\|202 | ···················· | ···················· | ····················· | ···················· | ···················· | ···················· | |
| K | 4413 | hCoV-19/Australia/VIC10290/2020\|EPI_ISL_562855\|202 | ···················· | ···················· | ····················· | ···················· | ···················· | ···················· | |
| K | 4414 | hCoV-19/Australia/VIC10292/2020\|EPI_ISL_562856\|202 | ···················· | ···················· | ····················· | ···················· | ···················· | ···················· | |
| K | 4415 | hCoV-19/Australia/VIC10294/2020\|EPI_ISL_562857\|202 | ···················· | ···················· | ····················· | ···················· | ···················· | ···················· | |
| K | 4416 | hCoV-19/Australia/VIC10296/2020\|EPI_ISL_562858\|202 | ···················· | ···················· | ····················· | ···················· | ···················· | ···················· | |
| K | 4417 | hCoV-19/Australia/VIC10299/2020\|EPI_ISL_562859\|202 | ···················· | ···················· | ····················· | ···················· | ···················· | ···················· | |
| K | 4419 | hCoV-19/Australia/VIC11504/2020\|EPI_ISL_563400\|202 | ···················· | ···················· | ····················· | ···················· | ···················· | ···················· | |

| | | | | | | | | | | |
|---|---|---|---|---|---|---|---|---|---|---|
| K | 4420 | hCoV-19/Australia/VIC11505/2020\|EPI_ISL_563401\|202 | ···················· | ···················· | ···················· | ···················· | ···················· | ···················· | | |
| K | 4421 | hCoV-19/Australia/VIC11511/2020\|EPI_ISL_563402\|202 | ···················· | ···················· | ···················· | ···················· | ···················· | ···················· | | |
| K | 4423 | hCoV-19/Australia/VIC11535/2020\|EPI_ISL_563404\|202 | ···················· | ···················· | ···················· | ···················· | ···················· | ···················· | | |
| K | 4424 | hCoV-19/Australia/VIC11536/2020\|EPI_ISL_563405\|202 | ···················· | ···················· | ···················· | ···················· | ···················· | ···················· | | |
| K | 4425 | hCoV-19/Australia/VIC11537/2020\|EPI_ISL_563406\|202 | ···················· | ···················· | ···················· | ···················· | ···················· | ···················· | | |
| K | 4426 | hCoV-19/Australia/VIC11545/2020\|EPI_ISL_563407\|202 | ···················· | ···················· | ···················· | ···················· | ···················· | ···················· | | |
| K | 4427 | hCoV-19/Australia/VIC11546/2020\|EPI_ISL_563408\|202 | ···················· | ···················· | ···················· | ···················· | ···················· | ···················· | | |
| K | 4430 | hCoV-19/Australia/VIC11550/2020\|EPI_ISL_563411\|202 | ···················· | ···················· | ···················· | ···················· | ···················· | ···················· | | |
| K | 4431 | hCoV-19/Australia/VIC11551/2020\|EPI_ISL_563412\|202 | ···················· | ···················· | ···················· | ···················· | ···················· | ···················· | | |
| K | 4432 | hCoV-19/Australia/VIC11552/2020\|EPI_ISL_563413\|202 | ···················· | ···················· | ···················· | ···················· | ···················· | ···················· | | |
| K | 4433 | hCoV-19/Australia/VIC11554/2020\|EPI_ISL_563414\|202 | ···················· | ···················· | ···················· | ···················· | ···················· | ···················· | | |
| K | 4435 | hCoV-19/Australia/VIC11565/2020\|EPI_ISL_563416\|202 | ···················· | ···················· | ···················· | ···················· | ···················· | ···················· | | |
| K | 4438 | hCoV-19/Australia/VIC11581/2020\|EPI_ISL_563419\|202 | ···················· | ···················· | ···················· | ···················· | ···················· | ···················· | | |
| K | 4439 | hCoV-19/Australia/VIC11582/2020\|EPI_ISL_563420\|202 | ···················· | ···················· | ···················· | ···················· | ···················· | ···················· | | |
| K | 4443 | hCoV-19/Australia/VIC11595/2020\|EPI_ISL_563424\|202 | ···················· | ···················· | ···················· | ···················· | ···················· | ···················· | | |
| K | 4444 | hCoV-19/Australia/VIC11596/2020\|EPI_ISL_563425\|202 | ···················· | ···················· | ···················· | ···················· | ···················· | ···················· | | |
| K | 4445 | hCoV-19/Australia/VIC11597/2020\|EPI_ISL_563426\|202 | ···················· | ···················· | ···················· | ···················· | ···················· | ···················· | | |
| K | 4446 | hCoV-19/Australia/VIC11598/2020\|EPI_ISL_563427\|202 | ···················· | ···················· | ·················R·· | ···················· | ···················· | ···················· | 1 | |
| K | 4447 | hCoV-19/Australia/VIC11600/2020\|EPI_ISL_563428\|202 | ···················· | ···················· | ···················· | ···················· | ···················· | ···················· | | |
| K | 4448 | hCoV-19/Australia/VIC11602/2020\|EPI_ISL_563429\|202 | ···················· | ···················· | ···················· | ···················· | ···················· | ···················· | | |
| K | 4449 | hCoV-19/Australia/VIC11607/2020\|EPI_ISL_563430\|202 | ···················· | ···················· | ···················· | ···················· | NNN················· | ···················· | | 1 |
| K | 4451 | hCoV-19/Australia/VIC11613/2020\|EPI_ISL_563432\|202 | ···················· | ···················· | ···················· | ···················· | ···················· | ···················· | | |
| K | 4452 | hCoV-19/Australia/VIC11683/2020\|EPI_ISL_563460\|202 | ···················· | ···················· | ···················· | ···················· | ···················· | ···················· | | |
| K | 4453 | hCoV-19/Australia/VIC11690/2020\|EPI_ISL_563461\|202 | ···················· | ···················· | ···················· | ···················· | ···················· | ···················· | | |
| K | 4454 | hCoV-19/Australia/VIC11697/2020\|EPI_ISL_563463\|202 | ···················· | ···················· | ···················· | ···················· | ···················· | ···················· | | |
| K | 4455 | hCoV-19/Australia/VIC11700/2020\|EPI_ISL_563464\|202 | ···················· | ···················· | ···················· | ···················· | ···················· | ···················· | | |
| K | 4457 | hCoV-19/Australia/VIC11739/2020\|EPI_ISL_563477\|202 | ···················· | ···················· | ···················· | ···················· | ···················· | ···················· | | |
| K | 4458 | hCoV-19/Australia/VIC11740/2020\|EPI_ISL_563478\|202 | ···················· | ···················· | ···················· | ···················· | ···················· | ···················· | | |
| K | 4459 | hCoV-19/Australia/VIC11741/2020\|EPI_ISL_563479\|202 | ···················· | ···················· | ···················· | ···················· | ···················· | ···················· | | |
| K | 4460 | hCoV-19/Australia/VIC11743/2020\|EPI_ISL_563480\|202 | ···················· | ···················· | ···················· | ···················· | ···················· | ···················· | | |
| K | 4461 | hCoV-19/Australia/VIC11745/2020\|EPI_ISL_563481\|202 | ···················· | ···················· | ···················· | ···················· | ···················· | ···················· | | |
| K | 4462 | hCoV-19/Australia/VIC11747/2020\|EPI_ISL_563482\|202 | ···················· | ···················· | ···················· | ···················· | ···················· | ···················· | | |
| K | 4463 | hCoV-19/Australia/VIC11749/2020\|EPI_ISL_563483\|202 | ···················· | ···················· | ···················· | ···················· | ···················· | ···················· | | |
| K | 4464 | hCoV-19/Australia/VIC11753/2020\|EPI_ISL_563484\|202 | ···················· | ···················· | ···················· | ···················· | ···················· | ···················· | | |
| K | 4465 | hCoV-19/Australia/VIC11754/2020\|EPI_ISL_563485\|202 | ···················· | ···················· | ···················· | ···················· | ···················· | ···················· | | |
| K | 4466 | hCoV-19/Australia/VIC11756/2020\|EPI_ISL_563486\|202 | ···················· | ···················· | ···················· | ···················· | ···················· | ···················· | | |
| K | 4467 | hCoV-19/Australia/VIC11757/2020\|EPI_ISL_563487\|202 | ···················· | ···················· | ···················· | ···················· | ···················· | ···················· | | |
| K | 4468 | hCoV-19/Australia/VIC11759/2020\|EPI_ISL_563488\|202 | ···················· | ···················· | ···················· | ···················· | ···················· | ···················· | | |
| K | 4469 | hCoV-19/Australia/VIC11760/2020\|EPI_ISL_563489\|202 | ···················· | ···················· | ···················· | ···················· | ···················· | ···················· | | |
| K | 4470 | hCoV-19/Australia/VIC11761/2020\|EPI_ISL_563490\|202 | ···················· | ···················· | ···················· | ···················· | ···················· | ···················· | | |
| K | 4471 | hCoV-19/Australia/VIC11762/2020\|EPI_ISL_563491\|202 | ···················· | ···················· | ···················· | ···················· | ···················· | ···················· | | |
| K | 4472 | hCoV-19/Australia/VIC11763/2020\|EPI_ISL_563492\|202 | ···················· | ···················· | ···················· | ···················· | ···················· | ········R··········· | | |
| K | 4475 | hCoV-19/Australia/VIC11774/2020\|EPI_ISL_563495\|202 | ···················· | ···················· | ···················· | ···················· | ···················· | ···················· | | |
| K | 4476 | hCoV-19/Australia/VIC11778/2020\|EPI_ISL_563496\|202 | ···················· | ···················· | ···················· | ···················· | ···················· | ···················· | | |
| K | 4477 | hCoV-19/Australia/VIC11850/2020\|EPI_ISL_563497\|202 | ···················· | ···················· | ···················· | ···················· | ···················· | ···················· | | |
| K | 4478 | hCoV-19/Australia/VIC11927/2020\|EPI_ISL_563498\|202 | ···················· | ···················· | ···················· | ···················· | ···················· | ···················· | | |
| K | 4479 | hCoV-19/Australia/VIC11937/2020\|EPI_ISL_563499\|202 | ···················· | ···················· | ···················· | ···················· | ···················· | ···················· | | |
| K | 4480 | hCoV-19/Australia/VIC7849/2020\|EPI_ISL_564700\|2020 | ···················· | ···················· | ···················· | ···················· | ···················· | ···················· | | |
| K | 4481 | hCoV-19/Australia/VIC7850/2020\|EPI_ISL_564701\|2020 | ···················· | ···················· | ···················· | ···················· | ···················· | ···················· | | |
| K | 4482 | hCoV-19/Australia/VIC7851/2020\|EPI_ISL_564702\|2020 | ···················· | ···················· | ···················· | ···················· | ···················· | ···················· | | |
| K | 4483 | hCoV-19/Australia/VIC7852/2020\|EPI_ISL_564703\|2020 | ···················· | ···················· | ···················· | ···················· | ···················· | ···················· | | |
| K | 4484 | hCoV-19/Australia/VIC7853/2020\|EPI_ISL_564704\|2020 | ···················· | ···················· | ···················· | ···················· | ···················· | ···················· | | |
| K | 4485 | hCoV-19/Australia/VIC7854/2020\|EPI_ISL_564705\|2020 | ···················· | ···················· | ···················· | ···················· | ···················· | ···················· | | |
| K | 4486 | hCoV-19/Australia/VIC7855/2020\|EPI_ISL_564706\|2020 | ···················· | ···················· | ···················· | ···················· | ···················· | ···················· | | |
| K | 4487 | hCoV-19/Australia/VIC7856/2020\|EPI_ISL_564707\|2020 | ···················· | ···················· | ···················· | ···················· | ···················· | ···················· | | |
| K | 4488 | hCoV-19/Australia/VIC7857/2020\|EPI_ISL_564708\|2020 | ···················· | ···················· | ···················· | ···················· | ···················· | ···················· | | |
| K | 4489 | hCoV-19/Australia/VIC7859/2020\|EPI_ISL_564709\|2020 | ···················· | ···················· | ···················· | ···················· | ···················· | ···················· | | |
| K | 4490 | hCoV-19/Australia/VIC7860/2020\|EPI_ISL_564710\|2020 | ···················· | ···················· | ···················· | ···················· | ···················· | ···················· | | |
| K | 4491 | hCoV-19/Australia/VIC7861/2020\|EPI_ISL_564711\|2020 | ···················· | ···················· | ···················· | ···················· | ···················· | ···················· | | |
| K | 4492 | hCoV-19/Australia/VIC7862/2020\|EPI_ISL_564712\|2020 | ···················· | ···················· | ···················· | ···················· | ···················· | ···················· | | |
| K | 4493 | hCoV-19/Australia/VIC7863/2020\|EPI_ISL_564713\|2020 | ···················· | ···················· | ···················· | ···················· | ···················· | ···················· | | |
| K | 4494 | hCoV-19/Australia/VIC7864/2020\|EPI_ISL_564714\|2020 | ···················· | ···················· | ···················· | ···················· | ···················· | ···················· | | |
| K | 4495 | hCoV-19/Australia/VIC7865/2020\|EPI_ISL_564715\|2020 | ···················· | ···················· | ···················· | ···················· | ···················· | ···················· | | |
| K | 4496 | hCoV-19/Australia/VIC7866/2020\|EPI_ISL_564716\|2020 | ···················· | ···················· | ···················· | ···················· | ···················· | ···················· | | |
| K | 4497 | hCoV-19/Australia/VIC7869/2020\|EPI_ISL_564717\|2020 | ···················· | ···················· | ···················· | ···················· | ···················· | ···················· | | |
| K | 4498 | hCoV-19/Australia/VIC7870/2020\|EPI_ISL_564718\|2020 | ···················· | ···················· | ···················· | ···················· | ···················· | ···················· | | |
| K | 4499 | hCoV-19/Australia/VIC7871/2020\|EPI_ISL_564719\|2020 | ···················· | ···················· | ···················· | ···················· | ···················· | ···················· | | |
| K | 4500 | hCoV-19/Australia/VIC7874/2020\|EPI_ISL_564720\|2020 | ···················· | ···················· | ···················· | ···················· | ···················· | ···················· | | |
| K | 4501 | hCoV-19/Australia/VIC7875/2020\|EPI_ISL_564721\|2020 | ···················· | ···················· | ···················· | ···················· | ···················· | ···················· | | |
| K | 4502 | hCoV-19/Australia/VIC7877/2020\|EPI_ISL_564722\|2020 | ···················· | ···················· | ···················· | ···················· | ···················· | ···················· | | |
| K | 4503 | hCoV-19/Australia/VIC7880/2020\|EPI_ISL_564723\|2020 | ···················· | ···················· | ···················· | ···················· | ···················· | ···················· | | |
| K | 4504 | hCoV-19/Australia/VIC7881/2020\|EPI_ISL_564724\|2020 | ···················· | ···················· | ···················· | ···················· | ···················· | ···················· | | |
| K | 4505 | hCoV-19/Australia/VIC7883/2020\|EPI_ISL_564725\|2020 | ···················· | ···················· | ···················· | ···················· | ···················· | ···················· | | |
| K | 4506 | hCoV-19/Australia/VIC7884/2020\|EPI_ISL_564726\|2020 | ···················· | ···················· | ···················· | ···················· | ···················· | ···················· | | |
| K | 4507 | hCoV-19/Australia/VIC7885/2020\|EPI_ISL_564727\|2020 | ···················· | ···················· | ···················· | ···················· | ···················· | ···················· | | |
| K | 4508 | hCoV-19/Australia/VIC7886/2020\|EPI_ISL_564728\|2020 | ···················· | ···················· | ···················· | ···················· | ···················· | ···················· | | |
| K | 4509 | hCoV-19/Australia/VIC7887/2020\|EPI_ISL_564729\|2020 | ···················· | ···················· | ···················· | ···················· | ···················· | ···················· | | |
| K | 4510 | hCoV-19/Australia/VIC7889/2020\|EPI_ISL_564730\|2020 | ···················· | ···················· | ···················· | ···················· | ···················· | ···················· | | |
| K | 4511 | hCoV-19/Australia/VIC7890/2020\|EPI_ISL_564731\|2020 | ···················· | ···················· | ···················· | ···················· | ···················· | ···················· | | |
| K | 4512 | hCoV-19/Australia/VIC7891/2020\|EPI_ISL_564732\|2020 | ···················· | ···················· | ···················· | ···················· | ···················· | ···················· | | |

| | | | | | | | | |
|---|---|---|---|---|---|---|---|---|
| K | 4513 | hCoV-19/Australia/VIC7894/2020\|EPI_ISL_564733\|2020 | ···················· | ···················· | ····················· | ···················· | ···················· | ···················· |
| K | 4514 | hCoV-19/Australia/VIC7895/2020\|EPI_ISL_564734\|2020 | ···················· | ···················· | ····················· | ···················· | ···················· | ···················· |
| K | 4515 | hCoV-19/Australia/VIC7897/2020\|EPI_ISL_564735\|2020 | ···················· | ···················· | ····················· | ···················· | ···················· | ···················· |
| K | 4516 | hCoV-19/Australia/VIC7899/2020\|EPI_ISL_564736\|2020 | ···················· | ···················· | ····················· | ···················· | ···················· | ···················· |
| K | 4517 | hCoV-19/Australia/VIC7903/2020\|EPI_ISL_564737\|2020 | ···················· | ···················· | ····················· | ···················· | ···················· | ···················· |
| K | 4518 | hCoV-19/Australia/VIC7908/2020\|EPI_ISL_564738\|2020 | ···················· | ···················· | ····················· | ···················· | ···················· | ···················· |
| K | 4519 | hCoV-19/Australia/VIC7910/2020\|EPI_ISL_564739\|2020 | ···················· | ···················· | ····················· | ···················· | ···················· | ···················· |
| K | 4520 | hCoV-19/Australia/VIC7911/2020\|EPI_ISL_564740\|2020 | ···················· | ···················· | ····················· | ···················· | ···················· | ···················· |
| K | 4521 | hCoV-19/Australia/VIC7912/2020\|EPI_ISL_564741\|2020 | ···················· | ···················· | ····················· | ···················· | ···················· | ···················· |
| K | 4522 | hCoV-19/Australia/VIC7916/2020\|EPI_ISL_564742\|2020 | ···················· | ···················· | ····················· | ···················· | ···················· | ···················· |
| K | 4523 | hCoV-19/Australia/VIC7920/2020\|EPI_ISL_564743\|2020 | ···················· | ···················· | ····················· | ···················· | ···················· | ···················· |
| K | 4524 | hCoV-19/Australia/VIC7921/2020\|EPI_ISL_564744\|2020 | ···················· | ···················· | ····················· | ···················· | ···················· | ···················· |
| K | 4525 | hCoV-19/Australia/VIC7923/2020\|EPI_ISL_564745\|2020 | ···················· | ···················· | ····················· | ···················· | ···················· | ···················· |
| K | 4526 | hCoV-19/Australia/VIC7924/2020\|EPI_ISL_564746\|2020 | ···················· | ···················· | ····················· | ···················· | ···················· | ···················· |
| K | 4527 | hCoV-19/Australia/VIC7925/2020\|EPI_ISL_564747\|2020 | ···················· | ···················· | ····················· | ···················· | ···K················ | ···················· |
| K | 4528 | hCoV-19/Australia/VIC7926/2020\|EPI_ISL_564748\|2020 | ···················· | ···················· | ····················· | ···················· | ···················· | ···················· |
| K | 4529 | hCoV-19/Australia/VIC7927/2020\|EPI_ISL_564749\|2020 | ···················· | ···················· | ····················· | ···················· | ···················· | ···················· |
| K | 4530 | hCoV-19/Australia/VIC7928/2020\|EPI_ISL_564750\|2020 | ···················· | ···················· | ····················· | ···················· | ···················· | ···················· |
| K | 4531 | hCoV-19/Australia/VIC7929/2020\|EPI_ISL_564751\|2020 | ···················· | ···················· | ····················· | ···················· | ···················· | ···················· |
| K | 4532 | hCoV-19/Australia/VIC7930/2020\|EPI_ISL_564752\|2020 | ···················· | ···················· | ····················· | ···················· | ···················· | ···················· |
| K | 4533 | hCoV-19/Australia/VIC7931/2020\|EPI_ISL_564753\|2020 | ···················· | ···················· | ····················· | ···················· | ···················· | ···················· |
| K | 4534 | hCoV-19/Australia/VIC7932/2020\|EPI_ISL_564754\|2020 | ···················· | ···················· | ····················· | ···················· | ···················· | ···················· |
| K | 4535 | hCoV-19/Australia/VIC7933/2020\|EPI_ISL_564755\|2020 | ···················· | ···················· | ····················· | ···················· | ···················· | ···················· |
| K | 4536 | hCoV-19/Australia/VIC7934/2020\|EPI_ISL_564756\|2020 | ···················· | ···················· | ····················· | ···················· | ···················· | ···················· |
| K | 4537 | hCoV-19/Australia/VIC7935/2020\|EPI_ISL_564757\|2020 | ···················· | ···················· | ····················· | ···················· | ···················· | ···················· |
| K | 4538 | hCoV-19/Australia/VIC7936/2020\|EPI_ISL_564758\|2020 | ···················· | ···················· | ····················· | ···················· | ···················· | ···················· |
| K | 4539 | hCoV-19/Australia/VIC7940/2020\|EPI_ISL_564759\|2020 | ···················· | ···················· | ····················· | ···················· | ···················· | ···················· |
| K | 4540 | hCoV-19/Australia/VIC7941/2020\|EPI_ISL_564760\|2020 | ···················· | ···················· | ····················· | ···················· | ···················· | ···················· |
| K | 4541 | hCoV-19/Australia/VIC7942/2020\|EPI_ISL_564761\|2020 | ···················· | ···················· | ····················· | ···················· | ···················· | ···················· |
| K | 4542 | hCoV-19/Australia/VIC7945/2020\|EPI_ISL_564762\|2020 | ···················· | ···················· | ····················· | ···················· | ···················· | ···················· |
| K | 4543 | hCoV-19/Australia/VIC7947/2020\|EPI_ISL_564763\|2020 | ···················· | ···················· | ····················· | ···················· | ···················· | ···················· |
| K | 4544 | hCoV-19/Australia/VIC7985/2020\|EPI_ISL_564790\|2020 | ···················· | ···················· | ····················· | ···················· | ···················· | ···················· |
| K | 4545 | hCoV-19/Australia/VIC7986/2020\|EPI_ISL_564791\|2020 | ···················· | ···················· | ····················· | ···················· | ···················· | ···················· |
| K | 4546 | hCoV-19/Australia/VIC7987/2020\|EPI_ISL_564792\|2020 | ···················· | ···················· | ····················· | ···················· | ···················· | ···················· |
| K | 4547 | hCoV-19/Australia/VIC7991/2020\|EPI_ISL_564794\|2020 | ···················· | ···················· | ····················· | ···················· | ···················· | ···················· |
| K | 4548 | hCoV-19/Australia/VIC7993/2020\|EPI_ISL_564795\|2020 | ···················· | ···················· | ····················· | ···················· | ···················· | ···················· |
| K | 4549 | hCoV-19/Australia/VIC7995/2020\|EPI_ISL_564796\|2020 | ···················· | ·····A·············· | ····················· | ···················· | ···················· | ···················· |
| K | 4550 | hCoV-19/Australia/VIC8508/2020\|EPI_ISL_565060\|2020 | ···················· | ···················· | ····················· | ···················· | ···················· | ···················· |
| K | 4551 | hCoV-19/Australia/VIC8509/2020\|EPI_ISL_565061\|2020 | ···················· | ···················· | ····················· | ···················· | ···················· | ···················· |
| K | 4552 | hCoV-19/Australia/VIC8510/2020\|EPI_ISL_565062\|2020 | ···················· | ···················· | ····················· | ···················· | ···················· | ···················· |
| K | 4553 | hCoV-19/Australia/VIC8511/2020\|EPI_ISL_565063\|2020 | ···················· | ···················· | ····················· | ···················· | ···················· | ···················· |
| K | 4554 | hCoV-19/Australia/VIC8513/2020\|EPI_ISL_565064\|2020 | ···················· | ···················· | ····················· | ···················· | ···················· | ···················· |
| K | 4555 | hCoV-19/Australia/VIC8514/2020\|EPI_ISL_565065\|2020 | ···················· | ···················· | ····················· | ···················· | ···················· | ···················· |
| K | 4556 | hCoV-19/Australia/VIC8515/2020\|EPI_ISL_565066\|2020 | ···················· | ···················· | ····················· | ···················· | ···················· | ···················· |
| K | 4557 | hCoV-19/Australia/VIC8517/2020\|EPI_ISL_565067\|2020 | ···················· | ···················· | ····················· | ···················· | ···················· | ···················· |
| K | 4558 | hCoV-19/Australia/VIC8518/2020\|EPI_ISL_565068\|2020 | ···················· | ···················· | ····················· | ···················· | ···················· | ···················· |
| K | 4559 | hCoV-19/Australia/VIC8519/2020\|EPI_ISL_565069\|2020 | ···················· | ···················· | ····················· | ···················· | ···················· | ···················· |
| K | 4560 | hCoV-19/Australia/VIC8520/2020\|EPI_ISL_565070\|2020 | ···················· | ···················· | ····················· | ···················· | ···················· | ···················· |
| K | 4561 | hCoV-19/Australia/VIC8521/2020\|EPI_ISL_565071\|2020 | ···················· | ···················· | ····················· | ···················· | ···················· | ···················· |
| K | 4562 | hCoV-19/Australia/VIC8523/2020\|EPI_ISL_565072\|2020 | ···················· | ···················· | ····················· | ···················· | ···················· | ···················· |
| K | 4563 | hCoV-19/Australia/VIC8524/2020\|EPI_ISL_565073\|2020 | ···················· | ···················· | ····················· | ···················· | ···················· | ···················· |
| K | 4564 | hCoV-19/Australia/VIC8525/2020\|EPI_ISL_565074\|2020 | ···················· | ···················· | ····················· | ···················· | ···················· | ···················· |
| K | 4565 | hCoV-19/Australia/VIC8526/2020\|EPI_ISL_565075\|2020 | ···················· | ···················· | ····················· | ···················· | ···················· | ···················· |
| K | 4566 | hCoV-19/Australia/VIC8527/2020\|EPI_ISL_565076\|2020 | ···················· | ···················· | ····················· | ···················· | ···················· | ···················· |
| K | 4567 | hCoV-19/Australia/VIC8528/2020\|EPI_ISL_565077\|2020 | ···················· | ···················· | ····················· | ···················· | ···················· | ···················· |
| K | 4568 | hCoV-19/Australia/VIC8530/2020\|EPI_ISL_565078\|2020 | ···················· | ···················· | ····················· | ···················· | ···················· | ···················· |
| K | 4569 | hCoV-19/Australia/VIC8531/2020\|EPI_ISL_565079\|2020 | ···················· | ···················· | ····················· | ···················· | ···················· | ···················· |
| K | 4570 | hCoV-19/Australia/VIC8532/2020\|EPI_ISL_565080\|2020 | ···················· | ···················· | ····················· | ···················· | ···················· | ···················· |
| K | 4571 | hCoV-19/Australia/VIC8536/2020\|EPI_ISL_565081\|2020 | ···················· | ···················· | ····················· | ···················· | ···················· | ···················· |
| K | 4572 | hCoV-19/Australia/VIC8541/2020\|EPI_ISL_565082\|2020 | ···················· | ···················· | ····················· | ···················· | ···················· | ···················· |
| K | 4573 | hCoV-19/Australia/VIC8545/2020\|EPI_ISL_565083\|2020 | ···················· | ···················· | ····················· | ···················· | ···················· | ···················· |
| K | 4574 | hCoV-19/Australia/VIC8547/2020\|EPI_ISL_565084\|2020 | ···················· | ···················· | ····················· | ···················· | ···················· | ···················· |
| K | 4575 | hCoV-19/Australia/VIC8548/2020\|EPI_ISL_565085\|2020 | ···················· | ···················· | ····················· | ···················· | ···················· | ···················· |
| K | 4576 | hCoV-19/Australia/VIC8549/2020\|EPI_ISL_565086\|2020 | ···················· | ···················· | ····················· | ···················· | ···················· | ···················· |
| K | 4577 | hCoV-19/Australia/VIC8550/2020\|EPI_ISL_565087\|2020 | ···················· | ···················· | ····················· | ···················· | ···················· | ···················· |
| K | 4578 | hCoV-19/Australia/VIC8552/2020\|EPI_ISL_565088\|2020 | ···················· | ···················· | ····················· | ···················· | ···················· | ···················· |
| K | 4579 | hCoV-19/Australia/VIC8558/2020\|EPI_ISL_565089\|2020 | ···················· | ···················· | ····················· | ···················· | ···················· | ···················· |
| K | 4580 | hCoV-19/Australia/VIC8559/2020\|EPI_ISL_565090\|2020 | ···················· | ···················· | ····················· | ···················· | ···················· | ···················· |
| K | 4581 | hCoV-19/Australia/VIC8562/2020\|EPI_ISL_565091\|2020 | ···················· | ···················· | ····················· | ···················· | ···················· | ···················· |
| K | 4582 | hCoV-19/Australia/VIC8563/2020\|EPI_ISL_565092\|2020 | ···················· | ···················· | ····················· | ···················· | ···················· | ···················· |
| K | 4583 | hCoV-19/Australia/VIC8565/2020\|EPI_ISL_565093\|2020 | ···················· | ···················· | ····················· | ···················· | ···················· | ···················· |
| K | 4584 | hCoV-19/Australia/VIC8566/2020\|EPI_ISL_565094\|2020 | ···················· | ···················· | ····················· | ···················· | ···················· | ···················· |
| K | 4585 | hCoV-19/Australia/VIC8568/2020\|EPI_ISL_565095\|2020 | ···················· | ···················· | ····················· | ···················· | ···················· | ···················· |
| K | 4587 | hCoV-19/Australia/VIC8571/2020\|EPI_ISL_565097\|2020 | ···················· | ···················· | ····················· | ···················· | ···················· | ···················· |
| K | 4588 | hCoV-19/Australia/VIC8577/2020\|EPI_ISL_565098\|2020 | ···················· | ···················· | ····················· | ···················· | ···················· | ···················· |
| K | 4589 | hCoV-19/Australia/VIC8580/2020\|EPI_ISL_565099\|2020 | ···················· | ···················· | ····················· | ···················· | ···················· | ···················· |
| K | 4590 | hCoV-19/England/ALDP-9CDABD/2020\|EPI_ISL_566390\|20 | ···················· | ···················· | ····················· | ···················· | ···················· | ···················· |
| K | 4591 | hCoV-19/England/ALDP-9CD6C8/2020\|EPI_ISL_566391\|20 | ···················· | ···················· | ····················· | ···················· | ···················· | ···················· |
| K | 4592 | hCoV-19/England/ALDP-9CDCE4/2020\|EPI_ISL_566392\|20 | ···················· | ···················· | ····················· | ···················· | ···················· | ···················· |
| K | 4593 | hCoV-19/England/ALDP-9CD09D/2020\|EPI_ISL_566393\|20 | ···················· | ···················· | ····················· | ···················· | ···················· | ···················· |

| | | | | | | | |
|---|---|---|---|---|---|---|---|
| K | 4594 hCoV-19/England/ALDP-9CD80E/2020\|EPI_ISL_566394\|20 | ···················· | ···················· | ···················· | ···················· | ···················· | ···················· |
| K | 4595 hCoV-19/England/ALDP-9CCF46/2020\|EPI_ISL_566395\|20 | ···················· | ·A·······A·········· | ···················· | ···················· | ···················· | ···················· |
| K | 4596 hCoV-19/England/ALDP-9CCF73/2020\|EPI_ISL_566396\|20 | ···················· | ···················· | ···················· | ···················· | ···················· | ···················· |
| K | 4597 hCoV-19/England/ALDP-9CCF55/2020\|EPI_ISL_566397\|20 | ···················· | ···················· | ···················· | ···················· | ···················· | ···················· |
| K | 4598 hCoV-19/England/ALDP-9CD6F5/2020\|EPI_ISL_566398\|20 | ···················· | ···················· | ···················· | ···················· | ···················· | ···················· |
| K | 4599 hCoV-19/England/ALDP-9CDF27/2020\|EPI_ISL_566399\|20 | ···················· | ···················· | ···················· | ···················· | ···················· | ···················· |
| K | 4600 hCoV-19/Scotland/QEUH-98F5A7/2020\|EPI_ISL_568291\|2 | ···················· | ···················· | ···················· | ···················· | ···················· | ···················· |
| K | 4601 hCoV-19/Scotland/QEUH-990673/2020\|EPI_ISL_568292\|2 | ···················· | ···················· | ···················· | ···················· | ···················· | ···················· |
| K | 4602 hCoV-19/Scotland/QEUH-99001B/2020\|EPI_ISL_568293\|2 | ···················· | ···················· | ···················· | ···················· | ···················· | ···················· |
| K | 4603 hCoV-19/Scotland/QEUH-98F9BA/2020\|EPI_ISL_568294\|2 | ···················· | ···················· | ···················· | ···················· | ···················· | ···················· |
| K | 4604 hCoV-19/Scotland/QEUH-9903C7/2020\|EPI_ISL_568295\|2 | ···················· | ···················· | ···················· | ···················· | ···················· | ···················· |
| K | 4605 hCoV-19/Scotland/QEUH-9903F4/2020\|EPI_ISL_568296\|2 | ···················· | ···················· | ···················· | ···················· | ···················· | ···················· |
| K | 4606 hCoV-19/Scotland/QEUH-98FE33/2020\|EPI_ISL_568297\|2 | ···················· | ···················· | ···················· | ···················· | ···················· | ···················· |
| K | 4607 hCoV-19/Scotland/QEUH-98F6D1/2020\|EPI_ISL_568298\|2 | ···················· | ···················· | ···················· | ···················· | ···················· | ···················· |
| K | 4608 hCoV-19/Scotland/QEUH-998D2B/2020\|EPI_ISL_568299\|2 | ···················· | ···················· | ···················· | ···················· | ···················· | ···················· |
| K | 4609 hCoV-19/Australia/VIC10251/2020\|EPI_ISL_562843\|202 | ···················· | ···················· | ···················· | ···················· | ···················· | ···················· |
| K | 4610 hCoV-19/Australia/VIC10253/2020\|EPI_ISL_562844\|202 | ···················· | ···················· | ···················· | ···················· | ···················· | ···················· |
| K | 4611 hCoV-19/Australia/VIC8420/2020\|EPI_ISL_565000\|2020 | ···················· | ···················· | ···················· | ···················· | ···················· | ···················· |
| K | 4612 hCoV-19/Australia/VIC8422/2020\|EPI_ISL_565001\|2020 | ···················· | ···················· | ···················· | ···················· | ···················· | ···················· |
| K | 4613 hCoV-19/Australia/VIC8424/2020\|EPI_ISL_565002\|2020 | ···················· | ···················· | ···················· | ···················· | ···················· | ···················· |
| K | 4614 hCoV-19/Australia/VIC8425/2020\|EPI_ISL_565003\|2020 | ···················· | ···················· | ···················· | ···················· | ···················· | ···················· |
| K | 4615 hCoV-19/Australia/VIC8427/2020\|EPI_ISL_565004\|2020 | ···················· | ···················· | ···················· | ···················· | ···················· | ···················· |
| K | 4616 hCoV-19/Australia/VIC8429/2020\|EPI_ISL_565005\|2020 | ···················· | ···················· | ···················· | ···················· | ···················· | ···················· |
| K | 4617 hCoV-19/Australia/VIC8431/2020\|EPI_ISL_565006\|2020 | ···················· | ···················· | ···················· | ···················· | ···················· | ···················· |
| K | 4618 hCoV-19/Australia/VIC8433/2020\|EPI_ISL_565007\|2020 | ···················· | ···················· | ···················· | ···················· | ···················· | ···················· |
| K | 4619 hCoV-19/Australia/VIC8434/2020\|EPI_ISL_565008\|2020 | ···················· | ···················· | ···················· | ···················· | ···················· | ···················· |
| K | 4620 hCoV-19/Australia/VIC8435/2020\|EPI_ISL_565009\|2020 | ···················· | ···················· | ···················· | ···················· | ···················· | ···················· |
| K | 4621 hCoV-19/Australia/VIC8436/2020\|EPI_ISL_565010\|2020 | ···················· | ···················· | ···················· | ···················· | ···················· | ···················· |
| K | 4622 hCoV-19/Australia/VIC8437/2020\|EPI_ISL_565011\|2020 | ···················· | ···················· | ···················· | ···················· | ···················· | ···················· |
| K | 4623 hCoV-19/Australia/VIC8438/2020\|EPI_ISL_565012\|2020 | ···················· | ···················· | ···················· | ···················· | ···················· | ···················· |
| K | 4624 hCoV-19/Australia/VIC8440/2020\|EPI_ISL_565013\|2020 | ···················· | ···················· | ···················· | ···················· | ···················· | ···················· |
| K | 4625 hCoV-19/Australia/VIC8441/2020\|EPI_ISL_565014\|2020 | ···················· | ···················· | ···················· | ···················· | ···················· | ···················· |
| K | 4626 hCoV-19/Australia/VIC8442/2020\|EPI_ISL_565015\|2020 | ···················· | ···················· | ···················· | ···················· | ···················· | ···················· |
| K | 4627 hCoV-19/Australia/VIC8443/2020\|EPI_ISL_565016\|2020 | ···················· | ···················· | ···················· | ···················· | ···················· | ···················· |
| K | 4628 hCoV-19/Australia/VIC8445/2020\|EPI_ISL_565017\|2020 | ···················· | ···················· | ···················· | ···················· | ···················· | ···················· |
| K | 4629 hCoV-19/Australia/VIC8446/2020\|EPI_ISL_565018\|2020 | ···················· | ···················· | ···················· | ···················· | ···················· | ···················· |
| K | 4630 hCoV-19/Australia/VIC8447/2020\|EPI_ISL_565019\|2020 | ···················· | ···················· | ···················· | ···················· | ···················· | ···················· |
| K | 4631 hCoV-19/Australia/VIC8448/2020\|EPI_ISL_565020\|2020 | ···················· | ···················· | ···················· | ···················· | ···················· | ···················· |
| K | 4632 hCoV-19/Australia/VIC8449/2020\|EPI_ISL_565021\|2020 | ···················· | ···················· | ···················· | ···················· | ···················· | ···················· |
| K | 4633 hCoV-19/Australia/VIC8450/2020\|EPI_ISL_565022\|2020 | ···················· | ···················· | ···················· | ···················· | ···················· | ···················· |
| K | 4634 hCoV-19/Australia/VIC8451/2020\|EPI_ISL_565023\|2020 | ···················· | ···················· | ···················· | ···················· | ···················· | ···················· |
| K | 4635 hCoV-19/Australia/VIC8452/2020\|EPI_ISL_565024\|2020 | ···················· | ···················· | ···················· | ···················· | ···················· | ···················· |
| K | 4636 hCoV-19/Australia/VIC8454/2020\|EPI_ISL_565025\|2020 | ···················· | ···················· | ···················· | ···················· | ···················· | ···················· |
| K | 4637 hCoV-19/Australia/VIC8455/2020\|EPI_ISL_565026\|2020 | ···················· | ···················· | ···················· | ···················· | ···················· | ···················· |
| K | 4638 hCoV-19/Australia/VIC8457/2020\|EPI_ISL_565027\|2020 | ···················· | ···················· | ···················· | ···················· | ···················· | ···················· |
| K | 4639 hCoV-19/Australia/VIC8458/2020\|EPI_ISL_565028\|2020 | ···················· | ···················· | ···················· | ···················· | ···················· | ···················· |
| K | 4640 hCoV-19/Australia/VIC8459/2020\|EPI_ISL_565029\|2020 | ···················· | ···················· | ···················· | ···················· | ···················· | ···················· |
| K | 4641 hCoV-19/Australia/VIC8460/2020\|EPI_ISL_565030\|2020 | ···················· | ···················· | ···················· | ···················· | ···················· | ···················· |
| K | 4642 hCoV-19/Australia/VIC8461/2020\|EPI_ISL_565031\|2020 | ···················· | ···················· | ···················· | ···················· | ···················· | ···················· |
| K | 4643 hCoV-19/Australia/VIC8462/2020\|EPI_ISL_565032\|2020 | ···················· | ···················· | ···················· | ···················· | ···················· | ···················· |
| K | 4644 hCoV-19/Australia/VIC8463/2020\|EPI_ISL_565033\|2020 | ···················· | ···················· | ···················· | ···················· | ···················· | ···················· |
| K | 4645 hCoV-19/Australia/VIC8464/2020\|EPI_ISL_565034\|2020 | ···················· | ···················· | ···················· | ···················· | ···················· | ···················· |
| K | 4646 hCoV-19/Australia/VIC8467/2020\|EPI_ISL_565035\|2020 | ···················· | ···················· | ···················· | ···················· | ···················· | ···················· |
| K | 4647 hCoV-19/Australia/VIC8468/2020\|EPI_ISL_565036\|2020 | ···················· | ···················· | ···················· | ···················· | ···················· | ···················· |
| K | 4648 hCoV-19/Australia/VIC8470/2020\|EPI_ISL_565037\|2020 | ···················· | ···················· | ···················· | ···················· | ···················· | ···················· |
| K | 4649 hCoV-19/Australia/VIC8471/2020\|EPI_ISL_565038\|2020 | ···················· | ···················· | ···················· | ···················· | ···················· | ···················· |
| K | 4650 hCoV-19/Australia/VIC8472/2020\|EPI_ISL_565039\|2020 | ···················· | ···················· | ···················· | ···················· | ···················· | ···················· |
| K | 4651 hCoV-19/Australia/VIC8474/2020\|EPI_ISL_565040\|2020 | ···················· | ···················· | ···················· | ···················· | ···················· | ···················· |
| K | 4652 hCoV-19/Australia/VIC8475/2020\|EPI_ISL_565041\|2020 | ···················· | ···················· | ···················· | ···················· | ···················· | ···················· |
| K | 4653 hCoV-19/Australia/VIC8477/2020\|EPI_ISL_565042\|2020 | ···················· | ···················· | ···················· | ···················· | ···················· | ···················· |
| K | 4654 hCoV-19/Australia/VIC8478/2020\|EPI_ISL_565043\|2020 | ···················· | ···················· | ···················· | ···················· | ···················· | ···················· |
| K | 4655 hCoV-19/Australia/VIC8479/2020\|EPI_ISL_565044\|2020 | ···················· | ···················· | ···················· | ···················· | ···················· | ···················· |
| K | 4656 hCoV-19/Australia/VIC8480/2020\|EPI_ISL_565045\|2020 | ···················· | ···················· | ···················· | ···················· | ···················· | ···················· |
| K | 4657 hCoV-19/Australia/VIC8481/2020\|EPI_ISL_565046\|2020 | ···················· | ···················· | ···················· | ···················· | ···················· | ···················· |
| K | 4658 hCoV-19/Australia/VIC8485/2020\|EPI_ISL_565047\|2020 | ···················· | ···················· | ···················· | ···················· | ···················· | ···················· |
| K | 4659 hCoV-19/Australia/VIC8486/2020\|EPI_ISL_565048\|2020 | ···················· | ···················· | ···················· | ···················· | ···················· | ···················· |
| K | 4660 hCoV-19/Australia/VIC8487/2020\|EPI_ISL_565049\|2020 | ···················· | ···················· | ···················· | ···················· | ···················· | ···················· |
| K | 4662 hCoV-19/Australia/VIC8495/2020\|EPI_ISL_565051\|2020 | ···················· | ···················· | ···················· | ···················· | ···················· | ···················· |
| K | 4663 hCoV-19/Australia/VIC8496/2020\|EPI_ISL_565052\|2020 | ···················· | ···················· | ···················· | ···················· | ···················· | ···················· |
| K | 4664 hCoV-19/Australia/VIC8497/2020\|EPI_ISL_565053\|2020 | ···················· | ···················· | ···················· | ···················· | ···················· | ···················· |
| K | 4665 hCoV-19/Australia/VIC8498/2020\|EPI_ISL_565054\|2020 | ···················· | ···················· | ···················· | ···················· | ···················· | ···················· |
| K | 4666 hCoV-19/Australia/VIC8501/2020\|EPI_ISL_565055\|2020 | ···················· | ···················· | ···················· | ···················· | ···················· | ···················· |
| K | 4667 hCoV-19/Australia/VIC8502/2020\|EPI_ISL_565056\|2020 | ···················· | ···················· | ···················· | ···················· | ···················· | ···················· |
| K | 4668 hCoV-19/Australia/VIC8505/2020\|EPI_ISL_565057\|2020 | ···················· | ···················· | ···················· | ···················· | ···················· | ···················· |
| K | 4669 hCoV-19/Australia/VIC8506/2020\|EPI_ISL_565058\|2020 | ···················· | ······A············· | ···················· | ···················· | ···················· | ···················· |
| K | 4670 hCoV-19/Australia/VIC8507/2020\|EPI_ISL_565059\|2020 | ···················· | ······A············· | ···················· | ···················· | ···················· | ···················· |
| K | 4671 hCoV-19/England/ALDP-9CE0AB/2020\|EPI_ISL_566304\|20 | ···················· | ···················· | ···················· | ···················· | ···················· | ···················· |
| K | 4672 hCoV-19/England/ALDP-9CD67D/2020\|EPI_ISL_566306\|20 | ···················· | ···················· | ···················· | ···················· | ···················· | ···················· |
| K | 4673 hCoV-19/England/ALDP-9CCF91/2020\|EPI_ISL_566308\|20 | ···················· | ·A·······A·········· | ···················· | ···················· | ···················· | ···················· |
| K | 4674 hCoV-19/England/ALDP-9CD0F7/2020\|EPI_ISL_566309\|20 | ···················· | ·A·················· | ···················· | ···················· | ···················· | ···A················ |

| | | | | | | | | |
|---|---|---|---|---|---|---|---|---|
| K | 4675 | hCoV-19/England/ALDP-9CD7F2/2020\|EPI_ISL_566310\|20 | ···················· | ···················· | ···················· | ···················· | ···················· | ···················· |
| K | 4676 | hCoV-19/England/ALDP-9CE102/2020\|EPI_ISL_566311\|20 | ···················· | ···················· | ···················· | ···················· | ···················· | ···················· |
| K | 4677 | hCoV-19/England/ALDP-9CDF90/2020\|EPI_ISL_566312\|20 | ···················· | ···················· | ···················· | ···················· | ···················· | ···················· |
| K | 4678 | hCoV-19/England/ALDP-9CD455/2020\|EPI_ISL_566313\|20 | ···················· | ·A·················· | ···················· | ···················· | ···················· | ···A················ |
| K | 4679 | hCoV-19/England/ALDP-9CDD1E/2020\|EPI_ISL_566314\|20 | ···················· | ···················· | ···················· | ···················· | ···················· | ···················· |
| K | 4680 | hCoV-19/England/ALDP-9CD7E3/2020\|EPI_ISL_566316\|20 | ···················· | ···················· | ···················· | ···················· | ···················· | ···················· |
| K | 4681 | hCoV-19/England/ALDP-9CCF64/2020\|EPI_ISL_566317\|20 | ···················· | ···················· | ···················· | ···················· | ···················· | ···················· |
| K | 4682 | hCoV-19/England/ALDP-9CD14F/2020\|EPI_ISL_566318\|20 | ···················· | ···················· | ···················· | ···················· | ····T··············· | ···················· |
| K | 4683 | hCoV-19/England/ALDP-9CCD6A/2020\|EPI_ISL_566319\|20 | ···················· | ···················· | ···················· | ···················· | ···················· | ···················· |
| K | 4684 | hCoV-19/England/ALDP-9CDF36/2020\|EPI_ISL_566320\|20 | ···················· | ···················· | ···················· | ···················· | ···················· | ···················· |
| K | 4685 | hCoV-19/England/ALDP-9CD7D4/2020\|EPI_ISL_566321\|20 | ···················· | ···················· | ···················· | ···················· | ···················· | ···················· |
| K | 4686 | hCoV-19/England/ALDP-9CCEA3/2020\|EPI_ISL_566322\|20 | ···················· | ···················· | ···················· | ···················· | ····T··············· | ···················· |
| K | 4687 | hCoV-19/England/ALDP-9CDA80/2020\|EPI_ISL_566323\|20 | ···················· | ···················· | ···················· | ···················· | ···················· | ···················· |
| K | 4688 | hCoV-19/England/ALDP-9CD15E/2020\|EPI_ISL_566325\|20 | ···················· | ···················· | ···················· | ···················· | ···················· | ···················· |
| K | 4689 | hCoV-19/England/ALDP-9CD507/2020\|EPI_ISL_566326\|20 | ···················· | ···················· | ···················· | ···················· | ···················· | ···················· |
| K | 4690 | hCoV-19/England/ALDP-9CCFDD/2020\|EPI_ISL_566327\|20 | ···················· | ···················· | ···················· | ···················· | ···················· | ···················· |
| K | 4691 | hCoV-19/England/ALDP-9CD65F/2020\|EPI_ISL_566328\|20 | ···················· | ···················· | ···················· | ···················· | ···················· | ···················· |
| K | 4692 | hCoV-19/England/ALDP-9CDBAB/2020\|EPI_ISL_566329\|20 | ···················· | ···················· | ···················· | ···················· | ···················· | ···················· |
| K | 4693 | hCoV-19/England/ALDP-9CD376/2020\|EPI_ISL_566330\|20 | ···················· | ···················· | ···················· | ···················· | ···················· | ···················· |
| K | 4694 | hCoV-19/England/ALDP-9CD859/2020\|EPI_ISL_566331\|20 | ···················· | ···················· | ···················· | ···················· | ···················· | ···················· |
| K | 4695 | hCoV-19/England/ALDP-9CD9FC/2020\|EPI_ISL_566333\|20 | ···················· | ···················· | ···················· | ···················· | ···················· | ···················· |
| K | 4696 | hCoV-19/England/ALDP-9CDDA5/2020\|EPI_ISL_566334\|20 | ···················· | ···················· | ···················· | ···················· | ···················· | ···················· |
| K | 4697 | hCoV-19/England/ALDP-9CCFA0/2020\|EPI_ISL_566335\|20 | ···················· | ···················· | ···················· | ···················· | ···················· | ···················· |
| K | 4698 | hCoV-19/England/ALDP-9CDD96/2020\|EPI_ISL_566336\|20 | ···················· | ·A·················· | ···················· | ···················· | ···················· | ···················· |
| K | 4699 | hCoV-19/England/ALDP-9CDEA2/2020\|EPI_ISL_566337\|20 | ···················· | ···················· | ···················· | ···················· | ···················· | ···················· |
| K | 4700 | hCoV-19/England/ALDP-9CDAEA/2020\|EPI_ISL_566338\|20 | ···················· | ·A·······A·········· | ···················· | ···················· | ···················· | ···················· |
| K | 4701 | hCoV-19/England/ALDP-9CD543/2020\|EPI_ISL_566339\|20 | ···················· | ·A·················· | ···················· | ···················· | ···················· | ···A················ |
| K | 4702 | hCoV-19/England/ALDP-9CDF81/2020\|EPI_ISL_566340\|20 | ···················· | ···················· | ···················· | ···················· | ···················· | ···················· |
| K | 4703 | hCoV-19/England/ALDP-9CDC11/2020\|EPI_ISL_566341\|20 | ···················· | ···················· | ···················· | ···················· | ···················· | ···················· |
| K | 4704 | hCoV-19/England/ALDP-9CDEFD/2020\|EPI_ISL_566342\|20 | ···················· | ···················· | ···················· | ···················· | ···················· | ···················· |
| K | 4705 | hCoV-19/England/ALDP-9CD051/2020\|EPI_ISL_566343\|20 | ···················· | ···················· | ···················· | ···················· | ···················· | ···················· |
| K | 4706 | hCoV-19/England/ALDP-9CCFFB/2020\|EPI_ISL_566345\|20 | ···················· | ···················· | ···················· | ···················· | ···················· | ···················· |
| K | 4707 | hCoV-19/England/ALDP-9CDC5D/2020\|EPI_ISL_566346\|20 | ···················· | ···················· | ···················· | ···················· | ······T············· | ···················· |
| K | 4708 | hCoV-19/England/ALDP-9CD1A9/2020\|EPI_ISL_566347\|20 | ···················· | ···················· | ···················· | ···················· | ···················· | ···················· |
| K | 4709 | hCoV-19/England/ALDP-9CCE49/2020\|EPI_ISL_566348\|20 | ···················· | ···················· | ···················· | ···················· | ···················· | ···················· |
| K | 4710 | hCoV-19/England/ALDP-9CD23D/2020\|EPI_ISL_566349\|20 | ···················· | ···················· | ···················· | ···················· | ···················· | ···················· |
| K | 4711 | hCoV-19/England/ALDP-9CD8B3/2020\|EPI_ISL_566350\|20 | ···················· | ···················· | ···················· | ···················· | ···················· | ···················· |
| K | 4712 | hCoV-19/England/ALDP-9CDEEE/2020\|EPI_ISL_566351\|20 | ···················· | ···················· | ···················· | ···················· | ···················· | ···················· |
| K | 4713 | hCoV-19/England/ALDP-9CD983/2020\|EPI_ISL_566352\|20 | ···················· | ···················· | ···················· | ···················· | ···················· | ···················· |
| K | 4714 | hCoV-19/England/ALDP-9CD5F8/2020\|EPI_ISL_566353\|20 | ···················· | ···················· | ···················· | ···················· | ···················· | ···················· |
| K | 4715 | hCoV-19/England/ALDP-9CD4CE/2020\|EPI_ISL_566354\|20 | ···················· | ···················· | ···················· | ···················· | ···················· | ···················· |
| K | 4716 | hCoV-19/England/ALDP-9CDE0C/2020\|EPI_ISL_566355\|20 | ···················· | ···················· | ···················· | ···················· | ······T············· | ···················· |
| K | 4717 | hCoV-19/England/ALDP-9CD19A/2020\|EPI_ISL_566356\|20 | ···················· | ···················· | ···················· | ···················· | ···················· | ···················· |
| K | 4718 | hCoV-19/England/ALDP-9CD103/2020\|EPI_ISL_566357\|20 | ···················· | ···················· | ···················· | ···················· | ···················· | ···················· |
| K | 4719 | hCoV-19/England/ALDP-9CDA44/2020\|EPI_ISL_566358\|20 | ···················· | ···················· | ···················· | ···················· | ···················· | ···················· |
| K | 4720 | hCoV-19/England/ALDP-9CDACC/2020\|EPI_ISL_566359\|20 | ···················· | ···················· | ···················· | ···················· | ···················· | ···················· |
| K | 4721 | hCoV-19/England/ALDP-9CD82C/2020\|EPI_ISL_566360\|20 | ···················· | ·A·················· | ···················· | ···················· | ···················· | ···A················ |
| K | 4722 | hCoV-19/England/ALDP-9CD5DA/2020\|EPI_ISL_566361\|20 | ···················· | ···················· | ···················· | ···················· | ···················· | ···················· |
| K | 4723 | hCoV-19/England/ALDP-9CCD4C/2020\|EPI_ISL_566362\|20 | ···················· | ···················· | ···················· | ···················· | ···················· | ···················· |
| K | 4724 | hCoV-19/England/ALDP-9CDA17/2020\|EPI_ISL_566363\|20 | ···················· | ·A·················· | ···················· | ···················· | ···················· | ···A················ |
| K | 4725 | hCoV-19/England/ALDP-9CDE93/2020\|EPI_ISL_566364\|20 | ···················· | ···················· | ···················· | ···················· | ···················· | ···················· |
| K | 4726 | hCoV-19/England/ALDP-9CD3D0/2020\|EPI_ISL_566366\|20 | ···················· | ·A·······A·········· | ···················· | ···················· | ···················· | ···················· |
| K | 4727 | hCoV-19/England/ALDP-9CD886/2020\|EPI_ISL_566367\|20 | ···················· | ···················· | ···················· | ···················· | ···················· | ···················· |
| K | 4728 | hCoV-19/England/ALDP-9CDB9C/2020\|EPI_ISL_566368\|20 | ···················· | ···················· | ···················· | ···················· | ···················· | ···················· |
| K | 4729 | hCoV-19/England/ALDP-9CDDE1/2020\|EPI_ISL_566369\|20 | ···················· | ···················· | ···················· | ···················· | ···················· | ···················· |
| K | 4730 | hCoV-19/England/ALDP-9CDADB/2020\|EPI_ISL_566370\|20 | ···················· | ···················· | ···················· | ···················· | ···················· | ···················· |
| K | 4731 | hCoV-19/England/ALDP-9CD604/2020\|EPI_ISL_566372\|20 | ···················· | ·A·················· | ···················· | ···················· | ···················· | ···A················ |
| K | 4732 | hCoV-19/England/ALDP-9CD83B/2020\|EPI_ISL_566373\|20 | ···················· | ···················· | ···················· | ···················· | ···················· | ···················· |
| K | 4733 | hCoV-19/England/ALDP-9CD6E6/2020\|EPI_ISL_566374\|20 | ···················· | ···················· | ···················· | ···················· | ···················· | ···················· |
| K | 4734 | hCoV-19/England/ALDP-9CDFEB/2020\|EPI_ISL_566375\|20 | ···················· | ···················· | ···················· | ···················· | ···················· | ···················· |
| K | 4735 | hCoV-19/England/ALDP-9CD297/2020\|EPI_ISL_566376\|20 | ···················· | ·A·················· | ···················· | ···················· | ···················· | ···················· |
| K | 4736 | hCoV-19/England/ALDP-9CD7C5/2020\|EPI_ISL_566378\|20 | ···················· | ···················· | ···················· | ···················· | ···················· | ···················· |
| K | 4737 | hCoV-19/England/ALDP-9CCED0/2020\|EPI_ISL_566380\|20 | ···················· | ·A·······A·········· | ···················· | ···················· | ···················· | ···················· |
| K | 4738 | hCoV-19/England/ALDP-9CDB32/2020\|EPI_ISL_566381\|20 | ···················· | ···················· | ···················· | ···················· | ···················· | ···················· |
| K | 4739 | hCoV-19/England/ALDP-9CDA53/2020\|EPI_ISL_566382\|20 | ···················· | ···················· | ···················· | ···················· | ···················· | ···················· |
| K | 4740 | hCoV-19/England/ALDP-9CD4FB/2020\|EPI_ISL_566383\|20 | ···················· | ···················· | ···················· | ···················· | ···················· | ···················· |
| K | 4741 | hCoV-19/England/ALDP-9CD73E/2020\|EPI_ISL_566384\|20 | ···················· | ···················· | ···················· | ···················· | ···················· | ···················· |
| K | 4742 | hCoV-19/England/ALDP-9CE09C/2020\|EPI_ISL_566386\|20 | ···················· | ···················· | ···················· | ···················· | ···················· | ···················· |
| K | 4743 | hCoV-19/England/ALDP-9CE1A8/2020\|EPI_ISL_566387\|20 | ···················· | ···················· | ···················· | ···················· | ···················· | ···················· |
| K | 4744 | hCoV-19/England/ALDP-9CD877/2020\|EPI_ISL_566388\|20 | ···················· | ···················· | ···················· | ···················· | ···················· | ···················· |
| K | 4745 | hCoV-19/England/ALDP-9CD570/2020\|EPI_ISL_566389\|20 | ···················· | ···················· | ···················· | ···················· | ···················· | ···················· |
| K | 4746 | hCoV-19/England/ALDP-9CD798/2020\|EPI_ISL_566400\|20 | ···················· | ···················· | ···················· | ···················· | ···················· | ···················· |
| K | 4747 | hCoV-19/England/ALDP-9CDBD8/2020\|EPI_ISL_566402\|20 | ···················· | ···················· | ···················· | ···················· | ···················· | ···················· |
| K | 4748 | hCoV-19/England/ALDP-9C5DCB/2020\|EPI_ISL_567635\|20 | ···················· | ···················· | ···················· | ···················· | ···················· | ···················· |
| K | 4749 | hCoV-19/England/ALDP-9C610A/2020\|EPI_ISL_567636\|20 | ···················· | ·A·················· | ···················· | ···················· | ···················· | ···A················ |
| K | 4750 | hCoV-19/England/ALDP-9C58E8/2020\|EPI_ISL_567637\|20 | ···················· | ···················· | ···················· | ···················· | ···················· | ···················· |
| K | 4751 | hCoV-19/England/ALDP-9C63F5/2020\|EPI_ISL_567638\|20 | ···················· | ···················· | ···················· | ···················· | ···················· | ···················· |
| K | 4752 | hCoV-19/England/ALDP-9C5FB6/2020\|EPI_ISL_567639\|20 | ···················· | ·A·················· | ···················· | ···················· | ···················· | ···A················ |
| K | 4753 | hCoV-19/England/ALDP-9C5A6A/2020\|EPI_ISL_567640\|20 | ···················· | ···················· | ···················· | ···················· | ···················· | ···················· |
| K | 4754 | hCoV-19/England/ALDP-9C5E40/2020\|EPI_ISL_567641\|20 | ···················· | ···················· | ···················· | ···················· | ···················· | ···················· |

| | | | | | | | | |
|---|---|---|---|---|---|---|---|---|
| K | 4755 | hCoV-19/England/ALDP-9C6410/2020\|EPI_ISL_567642\|20 | ···················· | ···················· | ···················· | ···················· | ···················· | ···················· |
| K | 4756 | hCoV-19/England/ALDP-9C50A4/2020\|EPI_ISL_567643\|20 | ···················· | ···················· | ···················· | ···················· | ···················· | ···················· |
| K | 4757 | hCoV-19/England/ALDP-9C5AE2/2020\|EPI_ISL_567644\|20 | ···················· | ···················· | ···················· | ···················· | ···················· | ···················· |
| K | 4758 | hCoV-19/England/ALDP-9C5833/2020\|EPI_ISL_567645\|20 | ···················· | ···················· | ···················· | ···················· | ···················· | ···················· |
| K | 4759 | hCoV-19/England/ALDP-9C5217/2020\|EPI_ISL_567646\|20 | ···················· | ···················· | ···················· | ···················· | ···················· | ···················· |
| K | 4760 | hCoV-19/England/ALDP-9C5CA0/2020\|EPI_ISL_567647\|20 | ···················· | ···················· | ···················· | ···················· | ···················· | ···················· |
| K | 4761 | hCoV-19/England/ALDP-9C504A/2020\|EPI_ISL_567648\|20 | ···················· | ···················· | ···················· | ···················· | ···················· | ···················· |
| K | 4762 | hCoV-19/England/ALDP-9C5D8F/2020\|EPI_ISL_567650\|20 | ···················· | ···················· | ···················· | ···················· | ···················· | ···················· |
| K | 4763 | hCoV-19/England/ALDP-9C5235/2020\|EPI_ISL_567651\|20 | ···················· | ···················· | ···················· | ···················· | ···················· | ···················· |
| K | 4764 | hCoV-19/England/ALDP-9C4E41/2020\|EPI_ISL_567653\|20 | ···················· | ···················· | ···················· | ···················· | ···················· | ···················· |
| K | 4765 | hCoV-19/England/ALDP-9C5BB2/2020\|EPI_ISL_567654\|20 | ···················· | ···················· | ···················· | ···················· | ···················· | ···················· |
| K | 4766 | hCoV-19/England/ALDP-9C5C64/2020\|EPI_ISL_567655\|20 | ···················· | ···················· | ···················· | ···················· | ···················· | ···················· |
| K | 4767 | hCoV-19/England/ALDP-9C5693/2020\|EPI_ISL_567656\|20 | ···················· | ···················· | ···················· | ···················· | ···················· | ···················· |
| K | 4768 | hCoV-19/England/ALDP-9C52BD/2020\|EPI_ISL_567657\|20 | ···················· | ·A·················· | ···················· | ···················· | ···················· | ···A················ |
| K | 4769 | hCoV-19/England/ALDP-9C529F/2020\|EPI_ISL_567658\|20 | ···················· | ·A·················· | ···················· | ···················· | ···················· | ···················· |
| K | 4770 | hCoV-19/England/ALDP-9C502C/2020\|EPI_ISL_567659\|20 | ···················· | ···················· | ···················· | ···················· | ···················· | ···················· |
| K | 4771 | hCoV-19/England/ALDP-9C5D61/2020\|EPI_ISL_567660\|20 | ···················· | ·A·······A·········· | ···················· | ···················· | ···················· | ···················· |
| K | 4772 | hCoV-19/England/ALDP-9C51CF/2020\|EPI_ISL_567661\|20 | ···················· | ···················· | ···················· | ···················· | ···················· | ···················· |
| K | 4773 | hCoV-19/England/ALDP-9C5EAA/2020\|EPI_ISL_567662\|20 | ···················· | ···················· | ···················· | ···················· | ···················· | ···················· |
| K | 4774 | hCoV-19/England/ALDP-9C5AC4/2020\|EPI_ISL_567663\|20 | ···················· | ·A·················· | ···················· | ···················· | ···················· | ···················· |
| K | 4775 | hCoV-19/England/ALDP-9C4B2C/2020\|EPI_ISL_567664\|20 | ···················· | ···················· | ···················· | ···················· | ···················· | ···················· |
| K | 4776 | hCoV-19/England/ALDP-9C628F/2020\|EPI_ISL_567665\|20 | ···················· | ·A·················· | ···················· | ···················· | ···················· | ···A················ |
| K | 4777 | hCoV-19/England/ALDP-9C4D26/2020\|EPI_ISL_567666\|20 | ···················· | ···················· | ···················· | ···················· | ···················· | ···················· |
| K | 4778 | hCoV-19/England/ALDP-9C5B3A/2020\|EPI_ISL_567667\|20 | ···················· | ···················· | ···················· | ···················· | ···················· | ···················· |
| K | 4779 | hCoV-19/England/ALDP-9C4F8A/2020\|EPI_ISL_567669\|20 | ···················· | ···················· | ···················· | ···················· | ···················· | ···················· |
| K | 4780 | hCoV-19/England/ALDP-9C5F3E/2020\|EPI_ISL_567670\|20 | ···················· | ···················· | ···················· | ···················· | ···················· | ···················· |
| K | 4781 | hCoV-19/England/ALDP-9C5C82/2020\|EPI_ISL_567671\|20 | ···················· | ···················· | ···················· | ···················· | ···················· | ···················· |
| K | 4782 | hCoV-19/England/ALDP-9C55F0/2020\|EPI_ISL_567672\|20 | ···················· | ·A·················· | ···················· | ···················· | ···················· | ···················· |
| K | 4783 | hCoV-19/England/ALDP-9C53AB/2020\|EPI_ISL_567673\|20 | ···················· | ···················· | ···················· | ···················· | ···················· | ···················· |
| K | 4784 | hCoV-19/England/ALDP-9C4D08/2020\|EPI_ISL_567674\|20 | ···················· | ···················· | ···················· | ···················· | ···················· | ···················· |
| K | 4785 | hCoV-19/England/ALDP-9C58D9/2020\|EPI_ISL_567675\|20 | ···················· | ···················· | ···················· | ···················· | ···················· | ···················· |
| K | 4786 | hCoV-19/England/ALDP-9C54F3/2020\|EPI_ISL_567676\|20 | ···················· | ···················· | ···················· | ···················· | ···················· | ···················· |
| K | 4787 | hCoV-19/England/ALDP-9C58F7/2020\|EPI_ISL_567677\|20 | ···················· | ···················· | ···················· | ···················· | ···················· | ···················· |
| K | 4788 | hCoV-19/England/ALDP-9C5F89/2020\|EPI_ISL_567680\|20 | ···················· | ···················· | ···················· | ···················· | ···················· | ···················· |
| K | 4789 | hCoV-19/England/ALDP-9C5A1F/2020\|EPI_ISL_567681\|20 | ···················· | ···················· | ···················· | ···················· | ···················· | ···················· |
| K | 4790 | hCoV-19/England/ALDP-9C5DF8/2020\|EPI_ISL_567682\|20 | ···················· | ···················· | ···················· | ···················· | ···················· | ···················· |
| K | 4791 | hCoV-19/England/ALDP-9C4FA8/2020\|EPI_ISL_567683\|20 | ···················· | ···················· | ···················· | ···················· | ···················· | ···················· |
| K | 4792 | hCoV-19/England/ALDP-9C4CA1/2020\|EPI_ISL_567684\|20 | ···················· | ···················· | ···················· | ···················· | ···················· | ···················· |
| K | 4793 | hCoV-19/England/ALDP-9C59B8/2020\|EPI_ISL_567685\|20 | ···················· | ···················· | ···················· | ···················· | ···················· | ···················· |
| K | 4794 | hCoV-19/England/ALDP-9C551E/2020\|EPI_ISL_567686\|20 | ···················· | ···················· | ···················· | ···················· | ···················· | ···················· |
| K | 4795 | hCoV-19/England/ALDP-9C4D80/2020\|EPI_ISL_567687\|20 | ···················· | ···················· | ···················· | ···················· | ···················· | ···················· |
| K | 4796 | hCoV-19/England/ALDP-9C511A/2020\|EPI_ISL_567688\|20 | ···················· | ···················· | ···················· | ···················· | ···················· | ···················· |
| K | 4797 | hCoV-19/England/ALDP-9C61EC/2020\|EPI_ISL_567689\|20 | ···················· | ···················· | ···················· | ···················· | ···················· | ···················· |
| K | 4798 | hCoV-19/England/ALDP-9C4CCF/2020\|EPI_ISL_567690\|20 | ···················· | ···················· | ···················· | ···················· | ···················· | ···················· |
| K | 4799 | hCoV-19/England/ALDP-9C5192/2020\|EPI_ISL_567691\|20 | ···················· | ···················· | ···················· | ···················· | ···················· | ···················· |
| K | 4800 | hCoV-19/England/ALDP-9C5666/2020\|EPI_ISL_567692\|20 | ···················· | ···················· | ···················· | ···················· | ···················· | ···················· |
| K | 4801 | hCoV-19/England/ALDP-9C4E32/2020\|EPI_ISL_567693\|20 | ···················· | ···················· | ···················· | ···················· | ···················· | ···················· |
| K | 4802 | hCoV-19/England/ALDP-9C537E/2020\|EPI_ISL_567694\|20 | ···················· | ···················· | ···················· | ···················· | ···················· | ···················· |
| K | 4803 | hCoV-19/England/ALDP-9C57AF/2020\|EPI_ISL_567695\|20 | ···················· | ···················· | ···················· | ···················· | ···················· | ···················· |
| K | 4804 | hCoV-19/England/ALDP-9C5912/2020\|EPI_ISL_567696\|20 | ···················· | ···················· | ···················· | ···················· | ···················· | ···················· |
| K | 4805 | hCoV-19/England/ALDP-9C4C83/2020\|EPI_ISL_567697\|20 | ···················· | ···················· | ···················· | ···················· | ···················· | ···················· |
| K | 4806 | hCoV-19/England/ALDP-9C588E/2020\|EPI_ISL_567698\|20 | ···················· | ···················· | ···················· | ···················· | ···················· | ···················· |
| K | 4807 | hCoV-19/England/ALDP-9C5B85/2020\|EPI_ISL_567700\|20 | ···················· | ···················· | ···················· | ···················· | ···················· | ···················· |
| K | 4808 | hCoV-19/England/ALDP-9C5C73/2020\|EPI_ISL_567701\|20 | ···················· | ·A·················· | ···················· | ···················· | ···················· | ···A················ |
| K | 4809 | hCoV-19/England/ALDP-9C4DCC/2020\|EPI_ISL_567702\|20 | ···················· | ···················· | ···················· | ···················· | ···················· | ···················· |
| K | 4810 | hCoV-19/England/ALDP-9C503B/2020\|EPI_ISL_567703\|20 | ···················· | ···················· | ···················· | ···················· | ···················· | ···················· |
| K | 4811 | hCoV-19/England/ALDP-9C5D70/2020\|EPI_ISL_567704\|20 | ···················· | ···················· | ···················· | ···················· | ···················· | ···················· |
| K | 4812 | hCoV-19/England/ALDP-9C5271/2020\|EPI_ISL_567705\|20 | ···················· | ···················· | ···················· | ···················· | ···················· | ···················· |
| K | 4813 | hCoV-19/England/ALDP-9C602B/2020\|EPI_ISL_567706\|20 | ···················· | ·A·················· | ···················· | ···················· | ···················· | ···A················ |
| K | 4814 | hCoV-19/England/ALDP-9C5CBF/2020\|EPI_ISL_567707\|20 | ···················· | ·A·················· | ···················· | ···················· | ···················· | ···A················ |
| K | 4815 | hCoV-19/England/ALDP-9C635F/2020\|EPI_ISL_567708\|20 | ···················· | ···················· | ···················· | ···················· | ···················· | ···················· |
| K | 4816 | hCoV-19/England/ALDP-9C50B3/2020\|EPI_ISL_567709\|20 | ···················· | ···················· | ···················· | ···················· | ····T··············· | ···················· |
| K | 4817 | hCoV-19/England/ALDP-9C4FE4/2020\|EPI_ISL_567710\|20 | ···················· | ···················· | ···················· | ···················· | ···················· | ···················· |
| K | 4818 | hCoV-19/England/ALDP-9C5A79/2020\|EPI_ISL_567711\|20 | ···················· | ···················· | ···················· | ···················· | ···················· | ···················· |
| K | 4819 | hCoV-19/England/ALDP-9C59F4/2020\|EPI_ISL_567712\|20 | ···················· | ···················· | ···················· | ···················· | ···················· | ···················· |
| K | 4820 | hCoV-19/England/ALDP-9C4BB3/2020\|EPI_ISL_567713\|20 | ···················· | ···················· | ···················· | ···················· | ···················· | ···················· |
| K | 4821 | hCoV-19/England/ALDP-9C5F4D/2020\|EPI_ISL_567714\|20 | ···················· | ···················· | ···················· | ···················· | ···················· | ···················· |
| K | 4822 | hCoV-19/England/ALDP-9C5639/2020\|EPI_ISL_567715\|20 | ···················· | ···················· | ···················· | ···················· | ···················· | ···················· |
| K | 4823 | hCoV-19/England/ALDP-9C5F10/2020\|EPI_ISL_567716\|20 | ···················· | ···················· | ···················· | ···················· | ···················· | ···················· |
| K | 4824 | hCoV-19/England/ALDP-9C5CDD/2020\|EPI_ISL_567717\|20 | ···················· | ···················· | ···················· | ···················· | ···················· | ···················· |
| K | 4825 | hCoV-19/England/ALDP-9C4B4A/2020\|EPI_ISL_567718\|20 | ···················· | ···················· | ···················· | ···················· | ···················· | ···················· |
| K | 4826 | hCoV-19/England/ALDP-9C5FE3/2020\|EPI_ISL_567721\|20 | ···················· | ······A············· | ···················· | ···················· | ···················· | ···················· |
| K | 4827 | hCoV-19/England/ALDP-9C6313/2020\|EPI_ISL_567723\|20 | ···················· | ···················· | ···················· | ···················· | ······T············· | ···················· |
| K | 4828 | hCoV-19/England/ALDP-9C5208/2020\|EPI_ISL_567724\|20 | ···················· | ···················· | ···················· | ···················· | ···················· | ···················· |
| K | 4829 | hCoV-19/England/ALDP-9C4DDB/2020\|EPI_ISL_567725\|20 | ···················· | ···················· | ···················· | ···················· | ···················· | ···················· |
| K | 4830 | hCoV-19/England/ALDP-9C5B49/2020\|EPI_ISL_567726\|20 | ···················· | ···················· | ···················· | ···················· | ···················· | ···················· |
| K | 4831 | hCoV-19/England/ALDP-9C4FD5/2020\|EPI_ISL_567727\|20 | ···················· | ···················· | ···················· | ···················· | ···················· | ···················· |
| K | 4832 | hCoV-19/England/ALDP-9C5B0D/2020\|EPI_ISL_567728\|20 | ···················· | ···················· | ···················· | ···················· | ···················· | ···················· |
| K | 4833 | hCoV-19/England/ALDP-9C5ED7/2020\|EPI_ISL_567729\|20 | ···················· | ···················· | ···················· | ···················· | ···················· | ···················· |
| K | 4834 | hCoV-19/England/ALDP-9C5851/2020\|EPI_ISL_567730\|20 | ···················· | ···················· | ···················· | ···················· | ···················· | ···················· |

| | | | | | | | |
|---|---|---|---|---|---|---|---|
| K | 4835 hCoV-19/England/ALDP-9C5C28/2020\|EPI_ISL_567731\|20 | ···················· | ·A·················· | ···················· | ···················· | ···················· | ···················· |
| K | 4836 hCoV-19/England/ALDP-9C4DAE/2020\|EPI_ISL_567732\|20 | ···················· | ···················· | ···················· | ···················· | ···················· | ···················· |
| K | 4837 hCoV-19/England/ALDP-9C6331/2020\|EPI_ISL_567733\|20 | ···················· | ···················· | ···················· | ···················· | ···················· | ···················· |
| K | 4838 hCoV-19/France/PAC-IHU-0791/2020\|EPI_ISL_568966\|20 | ···················· | ···················· | ···················· | ···················· | ···················· | ···················· |
| K | 4839 hCoV-19/France/PAC-IHU-0793/2020\|EPI_ISL_568967\|20 | ···················· | ···················· | ···················· | ···················· | ···················· | ···················· |
| K | 4840 hCoV-19/France/PAC-IHU-0796/2020\|EPI_ISL_568968\|20 | ···················· | ···················· | ···················· | ···················· | ···················· | ···················· |
| K | 4841 hCoV-19/France/PAC-IHU-0798/2020\|EPI_ISL_568969\|20 | ···················· | ···················· | ···················· | ···················· | ···················· | ···················· |
| K | 4842 hCoV-19/France/PAC-IHU-0799/2020\|EPI_ISL_568970\|20 | ···················· | ···················· | ···················· | ···················· | ···················· | ···················· |
| K | 4843 hCoV-19/France/PAC-IHU-0803/2020\|EPI_ISL_568971\|20 | ···················· | ···················· | ···················· | ···················· | ···················· | ···················· |
| K | 4844 hCoV-19/France/PAC-IHU-0808/2020\|EPI_ISL_568972\|20 | ···················· | ···················· | ···················· | ···················· | ···················· | ···················· |
| K | 4845 hCoV-19/France/PAC-IHU-0810/2020\|EPI_ISL_568973\|20 | ···················· | ···················· | ···················· | ···················· | ···················· | ···················· |
| K | 4846 hCoV-19/France/PAC-IHU-0811/2020\|EPI_ISL_568974\|20 | ···················· | ···················· | ···················· | ···················· | ···················· | ···················· |
| K | 4847 hCoV-19/France/PAC-IHU-0813/2020\|EPI_ISL_568975\|20 | ···················· | ···················· | ···················· | ···················· | ···················· | ···················· |
| K | 4848 hCoV-19/France/PAC-IHU-0814/2020\|EPI_ISL_568976\|20 | ···················· | ···················· | ···················· | ···················· | ···················· | ···················· |
| K | 4849 hCoV-19/France/PAC-IHU-0817/2020\|EPI_ISL_568977\|20 | ···················· | ···················· | ···················· | ···················· | ···················· | ···················· |
| K | 4850 hCoV-19/France/PAC-IHU-0820/2020\|EPI_ISL_568978\|20 | ···················· | ···················· | ···················· | ···················· | ···················· | ···················· |
| K | 4851 hCoV-19/France/PAC-IHU-0822/2020\|EPI_ISL_568979\|20 | ···················· | ···················· | ···················· | ···················· | ···················· | ···················· |
| K | 4852 hCoV-19/France/PAC-IHU-0823/2020\|EPI_ISL_568980\|20 | ···················· | ···················· | ···················· | ···················· | ···················· | ···················· |
| K | 4853 hCoV-19/France/PAC-IHU-0827/2020\|EPI_ISL_568981\|20 | ···················· | ···················· | ···················· | ···················· | ···················· | ···················· |
| K | 4854 hCoV-19/France/PAC-IHU-0828/2020\|EPI_ISL_568982\|20 | ···················· | ···················· | ···················· | ···················· | ···················· | ···················· |
| K | 4855 hCoV-19/France/PAC-IHU-0830/2020\|EPI_ISL_568983\|20 | ···················· | ···················· | ···················· | ···················· | ···················· | ···················· |
| K | 4856 hCoV-19/France/PAC-IHU-0833/2020\|EPI_ISL_568984\|20 | ···················· | ···················· | ···················· | ···················· | ···················· | ···················· |
| K | 4857 hCoV-19/France/PAC-IHU-0852/2020\|EPI_ISL_568986\|20 | ···················· | ···················· | ···················· | ···················· | ···················· | ···················· |
| K | 4858 hCoV-19/France/PAC-IHU-0923/2020\|EPI_ISL_568992\|20 | ···················· | ···················· | ···················· | ···················· | ···················· | ···················· |
| K | 4859 hCoV-19/France/PAC-IHU-0926/2020\|EPI_ISL_568993\|20 | ···················· | ···················· | ···················· | ···················· | ···················· | ···················· |
| K | 4860 hCoV-19/France/PAC-IHU-0928/2020\|EPI_ISL_568994\|20 | ···················· | ···················· | ···················· | ···················· | ···················· | ···················· |
| K | 4861 hCoV-19/France/PAC-IHU-0930/2020\|EPI_ISL_568995\|20 | ···················· | ···················· | ···················· | ···················· | ···················· | ···················· |
| K | 4862 hCoV-19/France/PAC-IHU-0933/2020\|EPI_ISL_568996\|20 | ···················· | ···················· | ···················· | ···················· | ···················· | ···················· |
| K | 4863 hCoV-19/France/PAC-IHU-0941/2020\|EPI_ISL_568997\|20 | ···················· | ···················· | ···················· | ···················· | ···················· | ···················· |
| K | 4864 hCoV-19/France/PAC-IHU-0942/2020\|EPI_ISL_568998\|20 | ···················· | ····A··············· | ···················· | ···················· | ···················· | ···················· |
| K | 4865 hCoV-19/Australia/VIC10943/2020\|EPI_ISL_562700\|202 | ···················· | ···················· | ···················· | ···················· | ···················· | ···················· |
| K | 4866 hCoV-19/Australia/VIC11208/2020\|EPI_ISL_562701\|202 | ···················· | ···················· | ···················· | ···················· | ···················· | ···················· |
| K | 4867 hCoV-19/Australia/VIC11540/2020\|EPI_ISL_562702\|202 | ···················· | ···················· | ···················· | ···················· | ···················· | ···················· |
| K | 4868 hCoV-19/Australia/VIC13059/2020\|EPI_ISL_562703\|202 | ···················· | ···················· | ···················· | ···················· | ···················· | ···················· |
| K | 4869 hCoV-19/Australia/VIC6885/2020\|EPI_ISL_562704\|2020 | ···················· | ···················· | ···················· | ···················· | ···················· | ···················· |
| K | 4870 hCoV-19/Australia/VIC7151/2020\|EPI_ISL_562705\|2020 | ···················· | ···················· | ···················· | ···················· | ···················· | ···················· |
| K | 4871 hCoV-19/Australia/VIC8235/2020\|EPI_ISL_562706\|2020 | ···················· | ···················· | ···················· | ···················· | ···················· | ···················· |
| K | 4872 hCoV-19/Australia/VIC8928/2020\|EPI_ISL_562707\|2020 | ···················· | ···················· | ···················· | ···················· | ···················· | ···················· |
| K | 4873 hCoV-19/Australia/VIC9176/2020\|EPI_ISL_562708\|2020 | ···················· | ···················· | ···················· | ···················· | ···················· | ···················· |
| K | 4874 hCoV-19/Australia/VIC10009/2020\|EPI_ISL_562728\|202 | ···················· | ···················· | ···················· | ···················· | ···················· | ···················· |
| K | 4875 hCoV-19/Australia/VIC10082/2020\|EPI_ISL_562729\|202 | ···················· | ···················· | ···················· | ···················· | ···················· | ···················· |
| K | 4876 hCoV-19/Australia/VIC10084/2020\|EPI_ISL_562730\|202 | ···················· | ···················· | ···················· | ···················· | ···················· | ···················· |
| K | 4877 hCoV-19/Australia/VIC10111/2020\|EPI_ISL_562731\|202 | ···················· | ···················· | ···················· | ···················· | ···················· | ···················· |
| K | 4878 hCoV-19/Australia/VIC10179/2020\|EPI_ISL_562732\|202 | ···················· | ···················· | ···················· | ···················· | ···················· | ···················· |
| K | 4880 hCoV-19/Australia/VIC10390/2020\|EPI_ISL_562734\|202 | ···················· | ···················· | ···················· | ···················· | ···················· | ···················· |
| K | 4881 hCoV-19/Australia/VIC10393/2020\|EPI_ISL_562735\|202 | ···················· | ···················· | ···················· | ···················· | ···················· | ···················· |
| K | 4882 hCoV-19/Australia/VIC11078/2020\|EPI_ISL_562736\|202 | ···················· | ···················· | ···················· | ···················· | ···················· | ···················· |
| K | 4883 hCoV-19/Australia/VIC11218/2020\|EPI_ISL_562737\|202 | ···················· | ···················· | ···················· | ···················· | ···················· | ···················· |
| K | 4884 hCoV-19/Australia/VIC11408/2020\|EPI_ISL_562738\|202 | ···················· | ···················· | ···················· | ···················· | ···················· | ···················· |
| K | 4885 hCoV-19/Australia/VIC11494/2020\|EPI_ISL_562739\|202 | ···················· | ···················· | ···················· | ···················· | ···················· | ···················· |
| K | 4886 hCoV-19/Australia/VIC10304/2020\|EPI_ISL_562861\|202 | ···················· | ···················· | ···················· | ···················· | ···················· | ···················· |
| K | 4887 hCoV-19/Australia/VIC10306/2020\|EPI_ISL_562862\|202 | ···················· | ···················· | ···················· | ···················· | ···················· | ···················· |
| K | 4888 hCoV-19/Australia/VIC10307/2020\|EPI_ISL_562863\|202 | ···················· | ···················· | ···················· | ···················· | ···················· | ···················· |
| K | 4889 hCoV-19/Australia/VIC10308/2020\|EPI_ISL_562864\|202 | ···················· | ···················· | ···················· | ···················· | ···················· | ···················· |
| K | 4890 hCoV-19/Australia/VIC10311/2020\|EPI_ISL_562865\|202 | ···················· | ···················· | ···················· | ···················· | ···················· | ···················· |
| K | 4891 hCoV-19/Australia/VIC10316/2020\|EPI_ISL_562866\|202 | ···················· | ···················· | ···················· | ···················· | ···················· | ···················· |
| K | 4892 hCoV-19/Australia/VIC10318/2020\|EPI_ISL_562867\|202 | ···················· | ···················· | ···················· | ···················· | ···················· | ···················· |
| K | 4893 hCoV-19/Australia/VIC10319/2020\|EPI_ISL_562868\|202 | ···················· | ···················· | ···················· | ···················· | ···················· | ···················· |
| K | 4895 hCoV-19/Australia/VIC10322/2020\|EPI_ISL_562870\|202 | ···················· | ···················· | ···················· | ···················· | ···················· | ···················· |
| K | 4896 hCoV-19/Australia/VIC10323/2020\|EPI_ISL_562871\|202 | ···················· | ···················· | ···················· | ···················· | ···················· | ···················· |
| K | 4897 hCoV-19/Australia/VIC10331/2020\|EPI_ISL_562872\|202 | ···················· | ···················· | ···················· | ···················· | ···················· | ···················· |
| K | 4898 hCoV-19/Australia/VIC10333/2020\|EPI_ISL_562873\|202 | ···················· | ···················· | ···················· | ···················· | ···················· | ···················· |
| K | 4899 hCoV-19/Australia/VIC10334/2020\|EPI_ISL_562874\|202 | ···················· | ···················· | ···················· | ···················· | ···················· | ···················· |
| K | 4900 hCoV-19/Australia/VIC10335/2020\|EPI_ISL_562875\|202 | ···················· | ···················· | ···················· | ···················· | ···················· | ···················· |
| K | 4903 hCoV-19/Australia/VIC10339/2020\|EPI_ISL_562878\|202 | ···················· | ······A············· | ···················· | ···················· | ···················· | ···················· |
| K | 4904 hCoV-19/Australia/VIC10342/2020\|EPI_ISL_562879\|202 | ···················· | ···················· | ···················· | ···················· | ···················· | ···················· |
| K | 4906 hCoV-19/Australia/VIC10344/2020\|EPI_ISL_562881\|202 | ···················· | ···················· | ···················· | ···················· | ···················· | ···················· |
| K | 4908 hCoV-19/Australia/VIC10351/2020\|EPI_ISL_562883\|202 | ···················· | ···················· | ···················· | ···················· | ···················· | ···················· |
| K | 4909 hCoV-19/Australia/VIC10353/2020\|EPI_ISL_562884\|202 | ···················· | ···················· | ···················· | ···················· | ···················· | ···················· |
| K | 4910 hCoV-19/Australia/VIC10354/2020\|EPI_ISL_562885\|202 | ···················· | ···················· | ···················· | ···················· | ···················· | ···················· |
| K | 4911 hCoV-19/Australia/VIC10356/2020\|EPI_ISL_562886\|202 | ···················· | ···················· | ···················· | ···················· | ···················· | ···················· |
| K | 4912 hCoV-19/Australia/VIC10357/2020\|EPI_ISL_562887\|202 | ···················· | ···················· | ···················· | ···················· | ···················· | ···················· |
| K | 4913 hCoV-19/Australia/VIC10359/2020\|EPI_ISL_562888\|202 | ···················· | ···················· | ···················· | ···················· | ···················· | ···················· |
| K | 4916 hCoV-19/Australia/VIC10363/2020\|EPI_ISL_562891\|202 | ···················· | ···················· | ···················· | ···················· | ···················· | ···················· |
| K | 4917 hCoV-19/Australia/VIC10373/2020\|EPI_ISL_562892\|202 | ···················· | ···················· | ···················· | ···················· | ···················· | ···················· |
| K | 4919 hCoV-19/Australia/VIC10380/2020\|EPI_ISL_562894\|202 | ···················· | ···················· | ···················· | ···················· | ···················· | ···················· |
| K | 4921 hCoV-19/Australia/VIC10388/2020\|EPI_ISL_562896\|202 | ···················· | ···················· | ···················· | ···················· | ···················· | ···················· |
| K | 4922 hCoV-19/Australia/VIC10389/2020\|EPI_ISL_562897\|202 | ···················· | ···················· | ···················· | ···················· | ···················· | ···················· |
| K | 4923 hCoV-19/Australia/VIC10394/2020\|EPI_ISL_562898\|202 | ···················· | ···················· | ···················· | ···················· | ···················· | ···················· |
| K | 4924 hCoV-19/Australia/VIC10397/2020\|EPI_ISL_562899\|202 | ···················· | ···················· | ···················· | ···················· | ···················· | ···················· |

| | | | | | | | | |
|---|---|---|---|---|---|---|---|---|
| K | 4925 | hCoV-19/England/ALDP-9CD3EF/2020\|EPI_ISL_566200\|20 | ···················· | ···················· | ···················· | ···················· | ···················· | ···················· |
| K | 4926 | hCoV-19/England/ALDP-9CDCF3/2020\|EPI_ISL_566202\|20 | ···················· | ···················· | ···················· | ···················· | ···················· | ···················· |
| K | 4927 | hCoV-19/England/ALDP-9CDE57/2020\|EPI_ISL_566203\|20 | ···················· | ···················· | ···················· | ···················· | ····T··············· | ···················· |
| K | 4928 | hCoV-19/England/ALDP-9CD042/2020\|EPI_ISL_566204\|20 | ···················· | ···················· | ···················· | ···················· | ···················· | ···················· |
| K | 4929 | hCoV-19/England/ALDP-9CD006/2020\|EPI_ISL_566205\|20 | ···················· | ···················· | ···················· | ···················· | ···················· | ···················· |
| K | 4930 | hCoV-19/England/ALDP-9CD1D6/2020\|EPI_ISL_566206\|20 | ···················· | ···················· | ···················· | ···················· | ···················· | ···················· |
| K | 4931 | hCoV-19/England/ALDP-9CCF28/2020\|EPI_ISL_566207\|20 | ···················· | ·A·······A·········· | ···················· | ···················· | ···················· | ···················· |
| K | 4932 | hCoV-19/England/ALDP-9CCEEF/2020\|EPI_ISL_566210\|20 | ···················· | ·A·······A·········· | ···················· | ···················· | ···················· | ···················· |
| K | 4933 | hCoV-19/England/ALDP-9CDEDF/2020\|EPI_ISL_566211\|20 | ···················· | ···················· | ···················· | ···················· | ···················· | ···················· |
| K | 4934 | hCoV-19/England/ALDP-9CDA62/2020\|EPI_ISL_566212\|20 | ···················· | ···················· | ···················· | ···················· | ···················· | ···················· |
| K | 4935 | hCoV-19/England/ALDP-9CE14E/2020\|EPI_ISL_566213\|20 | ···················· | ···················· | ···················· | ···················· | ···················· | ···················· |
| K | 4936 | hCoV-19/England/ALDP-9CD015/2020\|EPI_ISL_566214\|20 | ···················· | ···················· | ···················· | ···················· | ···················· | ···················· |
| K | 4937 | hCoV-19/England/ALDP-9CDC20/2020\|EPI_ISL_566215\|20 | ···················· | ···················· | ···················· | ···················· | ···················· | ···················· |
| K | 4938 | hCoV-19/England/ALDP-9CDB14/2020\|EPI_ISL_566216\|20 | ···················· | ···················· | ···················· | ···················· | ···················· | ···················· |
| K | 4939 | hCoV-19/England/ALDP-9CD419/2020\|EPI_ISL_566217\|20 | ···················· | ···················· | ···················· | ···················· | ···················· | ···················· |
| K | 4940 | hCoV-19/England/ALDP-9CDE2A/2020\|EPI_ISL_566218\|20 | ···················· | ···················· | ···················· | ···················· | ···················· | ···················· |
| K | 4941 | hCoV-19/Wales/ALDP-9CD367/2020\|EPI_ISL_566219\|2020 | ···················· | ·A·················· | ···················· | ···················· | ···················· | ···················· |
| K | 4942 | hCoV-19/England/ALDP-9CDE1B/2020\|EPI_ISL_566220\|20 | ···················· | ···················· | ···················· | ···················· | ···················· | ···················· |
| K | 4943 | hCoV-19/England/ALDP-9CDDD2/2020\|EPI_ISL_566221\|20 | ···················· | ···················· | ···················· | ···················· | ···················· | ···················· |
| K | 4944 | hCoV-19/England/ALDP-9CD200/2020\|EPI_ISL_566222\|20 | ···················· | ···················· | ···················· | ···················· | ···················· | ···················· |
| K | 4945 | hCoV-19/England/ALDP-9CD473/2020\|EPI_ISL_566223\|20 | ···················· | ···················· | ···················· | ···················· | ···················· | ···················· |
| K | 4946 | hCoV-19/England/ALDP-9CE18A/2020\|EPI_ISL_566224\|20 | ···················· | ···················· | ···················· | ···················· | ···················· | ···················· |
| K | 4947 | hCoV-19/England/ALDP-9CD8D1/2020\|EPI_ISL_566225\|20 | ···················· | ···················· | ···················· | ···················· | ···················· | ···················· |
| K | 4948 | hCoV-19/England/ALDP-9CCEB2/2020\|EPI_ISL_566226\|20 | ···················· | ·A·······A·········· | ···················· | ···················· | ···················· | ···················· |
| K | 4949 | hCoV-19/England/ALDP-9CDB8D/2020\|EPI_ISL_566227\|20 | ···················· | ···················· | ···················· | ···················· | ···················· | ···················· |
| K | 4950 | hCoV-19/England/ALDP-9CDC7B/2020\|EPI_ISL_566228\|20 | ···················· | ···················· | ···················· | ···················· | ···················· | ···················· |
| K | 4951 | hCoV-19/England/ALDP-9CD6B9/2020\|EPI_ISL_566229\|20 | ···················· | ···················· | ···················· | ···················· | ···················· | ···················· |
| K | 4952 | hCoV-19/England/ALDP-9CD17C/2020\|EPI_ISL_566230\|20 | ···················· | ···················· | ···················· | ···················· | ···················· | ···················· |
| K | 4953 | hCoV-19/England/ALDP-9CDE48/2020\|EPI_ISL_566231\|20 | ···················· | ···················· | ···················· | ···················· | ···················· | ···················· |
| K | 4954 | hCoV-19/England/ALDP-9CCFBF/2020\|EPI_ISL_566234\|20 | ···················· | ·A·······A·········· | ···················· | ···················· | ···················· | ···················· |
| K | 4955 | hCoV-19/England/ALDP-9CD2F1/2020\|EPI_ISL_566235\|20 | ···················· | ·A·················· | ···················· | ···················· | ···················· | ···················· |
| K | 4956 | hCoV-19/England/ALDP-9CD701/2020\|EPI_ISL_566236\|20 | ···················· | ···················· | ···················· | ···················· | ···················· | ···················· |
| K | 4957 | hCoV-19/England/ALDP-9CD349/2020\|EPI_ISL_566237\|20 | ···················· | ···················· | ···················· | ···················· | ···················· | ···················· |
| K | 4958 | hCoV-19/Wales/ALDP-9CCF0A/2020\|EPI_ISL_566238\|2020 | ···················· | ···················· | ···················· | ···················· | ···················· | ···················· |
| K | 4959 | hCoV-19/England/ALDP-9CD622/2020\|EPI_ISL_566239\|20 | ···················· | ···················· | ···················· | ···················· | ···················· | ···················· |
| K | 4960 | hCoV-19/England/ALDP-9CD30D/2020\|EPI_ISL_566240\|20 | ···················· | ···················· | ···················· | ···················· | ···················· | ···················· |
| K | 4961 | hCoV-19/England/ALDP-9CDE75/2020\|EPI_ISL_566241\|20 | ···················· | ·A·················· | ···················· | ···················· | ···················· | ···A················ |
| K | 4962 | hCoV-19/England/ALDP-9CD464/2020\|EPI_ISL_566243\|20 | ···················· | ···················· | ···················· | ···················· | ···················· | ···················· |
| K | 4963 | hCoV-19/England/ALDP-9CDD69/2020\|EPI_ISL_566244\|20 | ···················· | ···················· | ···················· | ···················· | ···················· | ···················· |
| K | 4964 | hCoV-19/England/ALDP-9CDEC0/2020\|EPI_ISL_566245\|20 | ···················· | ···················· | ···················· | ···················· | ···················· | ···················· |
| K | 4965 | hCoV-19/England/ALDP-9CD21F/2020\|EPI_ISL_566246\|20 | ···················· | ···················· | ···················· | ···················· | ···················· | ···················· |
| K | 4966 | hCoV-19/England/ALDP-9CD9B0/2020\|EPI_ISL_566247\|20 | ···················· | ···················· | ···················· | ···················· | ···················· | ···················· |
| K | 4967 | hCoV-19/England/ALDP-9CE023/2020\|EPI_ISL_566248\|20 | ···················· | ···················· | ···················· | ···················· | ···················· | ···················· |
| K | 4968 | hCoV-19/England/ALDP-9CD22E/2020\|EPI_ISL_566270\|20 | ···················· | ···················· | ···················· | ···················· | ···················· | ···················· |
| K | 4969 | hCoV-19/England/ALDP-9CE15D/2020\|EPI_ISL_566271\|20 | ···················· | ···················· | ···················· | ···················· | ···················· | ···················· |
| K | 4970 | hCoV-19/England/ALDP-9CDF54/2020\|EPI_ISL_566272\|20 | ···················· | ···················· | ···················· | ···················· | ···················· | ···················· |
| K | 4971 | hCoV-19/England/ALDP-9CDE66/2020\|EPI_ISL_566273\|20 | ···················· | ·A·················· | ···················· | ···················· | ···················· | ···A················ |
| K | 4972 | hCoV-19/England/ALDP-9CDA08/2020\|EPI_ISL_566275\|20 | ···················· | ···················· | ···················· | ···················· | ···················· | ···················· |
| K | 4973 | hCoV-19/England/ALDP-9CD1F4/2020\|EPI_ISL_566277\|20 | ···················· | ···················· | ···················· | ···················· | ···················· | ···················· |
| K | 4974 | hCoV-19/England/ALDP-9CDE84/2020\|EPI_ISL_566278\|20 | ···················· | ···················· | ···················· | ···················· | ···················· | ···················· |
| K | 4975 | hCoV-19/England/ALDP-9CD3B2/2020\|EPI_ISL_566279\|20 | ···················· | ···················· | ···················· | ···················· | ···················· | ···················· |
| K | 4976 | hCoV-19/England/ALDP-9CD75C/2020\|EPI_ISL_566280\|20 | ···················· | ·A·················· | ···················· | ···················· | ···················· | ···A················ |
| K | 4977 | hCoV-19/England/ALDP-9CD3A3/2020\|EPI_ISL_566281\|20 | ···················· | ···················· | ···················· | ···················· | ···················· | ···················· |
| K | 4978 | hCoV-19/England/ALDP-9CD4BF/2020\|EPI_ISL_566282\|20 | ···················· | ···················· | ···················· | ···················· | ···················· | ···················· |
| K | 4979 | hCoV-19/England/ALDP-9CDFAF/2020\|EPI_ISL_566283\|20 | ···················· | ···················· | ···················· | ···················· | ···················· | ···················· |
| K | 4980 | hCoV-19/England/ALDP-9CD8E0/2020\|EPI_ISL_566284\|20 | ···················· | ···················· | ···················· | ···················· | ···················· | ···················· |
| K | 4981 | hCoV-19/England/ALDP-9CCE3A/2020\|EPI_ISL_566285\|20 | ···················· | ···················· | ···················· | ···················· | ···················· | ···················· |
| K | 4982 | hCoV-19/England/ALDP-9CE032/2020\|EPI_ISL_566286\|20 | ···················· | ···················· | ···················· | ···················· | ···················· | ···················· |
| K | 4983 | hCoV-19/England/ALDP-9CE0D8/2020\|EPI_ISL_566287\|20 | ···················· | ···················· | ···················· | ···················· | ···················· | ···················· |
| K | 4984 | hCoV-19/England/ALDP-9CE0C9/2020\|EPI_ISL_566288\|20 | ···················· | ···················· | ···················· | ···················· | ···················· | ···················· |
| K | 4985 | hCoV-19/England/ALDP-9CDD87/2020\|EPI_ISL_566290\|20 | ···················· | ···················· | ···················· | ···················· | ···················· | ···················· |
| K | 4986 | hCoV-19/England/ALDP-9CDE39/2020\|EPI_ISL_566291\|20 | ···················· | ···················· | ···················· | ···················· | ···················· | ···················· |
| K | 4987 | hCoV-19/England/ALDP-9CCDC4/2020\|EPI_ISL_566292\|20 | ···················· | ·A·················· | ···················· | ···················· | ···················· | ···A················ |
| K | 4988 | hCoV-19/England/ALDP-9CE0BA/2020\|EPI_ISL_566294\|20 | ···················· | ···················· | ···················· | ···················· | ···················· | ···················· |
| K | 4989 | hCoV-19/England/ALDP-9CDF72/2020\|EPI_ISL_566295\|20 | ···················· | ···················· | ···················· | ···················· | ···················· | ···················· |
| K | 4990 | hCoV-19/England/ALDP-9CDD2D/2020\|EPI_ISL_566296\|20 | ···················· | ···················· | ···················· | ···················· | ······T············· | ···················· |
| K | 4991 | hCoV-19/England/ALDP-9CD74D/2020\|EPI_ISL_566297\|20 | ···················· | ···················· | ···················· | ···················· | ···················· | ···················· |
| K | 4992 | hCoV-19/England/ALDP-9CD974/2020\|EPI_ISL_566298\|20 | ···················· | ···················· | ···················· | ···················· | ···················· | ···················· |
| K | 4993 | hCoV-19/England/ALDP-9CCE76/2020\|EPI_ISL_566299\|20 | ···················· | ···················· | ···················· | ···················· | ···················· | ···················· |
| K | 4994 | hCoV-19/England/ALDP-9CD516/2020\|EPI_ISL_566403\|20 | ···················· | ···················· | ···················· | ···················· | ···················· | ···················· |
| K | 4995 | hCoV-19/England/ALDP-9CD8A4/2020\|EPI_ISL_566404\|20 | ···················· | ·A·················· | ···················· | ···················· | ···················· | ···A················ |
| K | 4996 | hCoV-19/England/ALDP-9CCF19/2020\|EPI_ISL_566405\|20 | ···················· | ···················· | ···················· | ···················· | ···················· | ···················· |
| K | 4997 | hCoV-19/England/ALDP-9CD112/2020\|EPI_ISL_566407\|20 | ···················· | ···················· | ···················· | ···················· | ···················· | ···················· |
| K | 4998 | hCoV-19/England/ALDP-9CD26A/2020\|EPI_ISL_566408\|20 | ···················· | ···················· | ···················· | ···················· | ···················· | ···················· |
| K | 4999 | hCoV-19/England/ALDP-9CDF63/2020\|EPI_ISL_566409\|20 | ···················· | ···················· | ···················· | ···················· | ···················· | ···················· |
| K | 5000 | hCoV-19/England/ALDP-9CD0BB/2020\|EPI_ISL_566410\|20 | ···················· | ·A·················· | ···················· | ···················· | ···················· | ···A················ |
| K | 5001 | hCoV-19/England/ALDP-9CD2E2/2020\|EPI_ISL_566411\|20 | ···················· | ·A·················· | ···················· | ···················· | ···················· | ···A················ |
| K | 5002 | hCoV-19/England/ALDP-9CDDB4/2020\|EPI_ISL_566412\|20 | ···················· | ···················· | ···················· | ···················· | ···················· | ···················· |
| K | 5003 | hCoV-19/England/ALDP-9CDB41/2020\|EPI_ISL_566413\|20 | ···················· | ···················· | ···················· | ···················· | ···················· | ···················· |
| K | 5004 | hCoV-19/England/ALDP-9CCE67/2020\|EPI_ISL_566414\|20 | ···················· | ···················· | ···················· | ···················· | ···················· | ···················· |

| | | | | | | | | |
|---|---|---|---|---|---|---|---|---|
| K | 5005 | hCoV-19/England/ALDP-9CD31C/2020\|EPI_ISL_566415\|20 | ···················· | ···················· | ···················· | ···················· | ···················· | ···················· |
| K | 5006 | hCoV-19/England/ALDP-9CDFCD/2020\|EPI_ISL_566416\|20 | ···················· | ···················· | ···················· | ···················· | ···················· | ···················· |
| K | 5007 | hCoV-19/England/ALDP-9CE17B/2020\|EPI_ISL_566417\|20 | ···················· | ···················· | ···················· | ···················· | ···················· | ···················· |
| K | 5008 | hCoV-19/England/ALDP-9CD288/2020\|EPI_ISL_566418\|20 | ···················· | ·A·················· | ···················· | ···················· | ···················· | ···················· |
| K | 5009 | hCoV-19/England/ALDP-9CCE58/2020\|EPI_ISL_566419\|20 | ···················· | ···················· | ···················· | ···················· | ···················· | ···················· |
| K | 5010 | hCoV-19/England/ALDP-9CD5CB/2020\|EPI_ISL_566420\|20 | ···················· | ···················· | ···················· | ···················· | ···················· | ···················· |
| K | 5011 | hCoV-19/England/MILK-9C20C5/2020\|EPI_ISL_566421\|20 | ···················· | ···················· | ···················· | ···················· | ···················· | ···················· |
| K | 5012 | hCoV-19/England/CAMC-9B25AB/2020\|EPI_ISL_566423\|20 | ···················· | ···················· | ···················· | ···················· | ···················· | ···················· |
| K | 5013 | hCoV-19/England/MILK-9C1F05/2020\|EPI_ISL_566424\|20 | ···················· | ···················· | ···················· | ···················· | ···················· | ···················· |
| K | 5014 | hCoV-19/England/QEUH-9CA4A3/2020\|EPI_ISL_567514\|20 | ···················· | ···················· | ···················· | ···················· | ···················· | ···················· |
| K | 5015 | hCoV-19/England/QEUH-9CA8D4/2020\|EPI_ISL_567515\|20 | ···················· | ···················· | ···················· | ···················· | ···················· | ···················· |
| K | 5016 | hCoV-19/England/QEUH-9CA79B/2020\|EPI_ISL_567517\|20 | ···················· | ···················· | ···················· | ···················· | ···················· | ···················· |
| K | 5017 | hCoV-19/England/QEUH-9C994B/2020\|EPI_ISL_567518\|20 | ···················· | ···················· | ···················· | ···················· | ···················· | ···················· |
| K | 5018 | hCoV-19/England/QEUH-9C937A/2020\|EPI_ISL_567520\|20 | ···················· | ···················· | ···················· | ···················· | ···················· | ···················· |
| K | 5019 | hCoV-19/England/QEUH-9C987B/2020\|EPI_ISL_567521\|20 | ···················· | ·A·················· | ···················· | ···················· | ···················· | ···A················ |
| K | 5020 | hCoV-19/England/QEUH-9CA151/2020\|EPI_ISL_567522\|20 | ···················· | ···················· | ···················· | ···················· | ···················· | ···················· |
| K | 5021 | hCoV-19/England/QEUH-9C9FEF/2020\|EPI_ISL_567523\|20 | ···················· | ···················· | ···················· | ···················· | ···················· | ···················· |
| K | 5022 | hCoV-19/England/QEUH-9C93F2/2020\|EPI_ISL_567524\|20 | ···················· | ···················· | ···················· | ···················· | ···················· | ···················· |
| K | 5023 | hCoV-19/England/QEUH-9CA7F5/2020\|EPI_ISL_567525\|20 | ···················· | ···················· | ···················· | ···················· | ···················· | ···················· |
| K | 5024 | hCoV-19/England/QEUH-9CA1E8/2020\|EPI_ISL_567526\|20 | ···················· | ···················· | ···················· | ···················· | ···················· | ···················· |
| K | 5025 | hCoV-19/England/QEUH-9C9F1C/2020\|EPI_ISL_567528\|20 | ···················· | ···················· | ···················· | ···················· | ···················· | ···················· |
| K | 5026 | hCoV-19/England/QEUH-9C98C6/2020\|EPI_ISL_567529\|20 | ···················· | ·A·················· | ···················· | ···················· | ···················· | ···A················ |
| K | 5027 | hCoV-19/England/QEUH-9C96EA/2020\|EPI_ISL_567530\|20 | ···················· | ···················· | ···················· | ···················· | ······T············· | ···················· |
| K | 5028 | hCoV-19/England/QEUH-9C9D7C/2020\|EPI_ISL_567531\|20 | ···················· | ···················· | ···················· | ···················· | ···················· | ···················· |
| K | 5029 | hCoV-19/England/QEUH-9CA616/2020\|EPI_ISL_567532\|20 | ···················· | ···················· | ···················· | ···················· | ···················· | ···················· |
| K | 5030 | hCoV-19/England/ALDP-9C50FF/2020\|EPI_ISL_567533\|20 | ···················· | ···················· | ···················· | ···················· | ···················· | ···················· |
| K | 5031 | hCoV-19/England/ALDP-9C5420/2020\|EPI_ISL_567534\|20 | ···················· | ···················· | ···················· | ···················· | ···················· | ···················· |
| K | 5032 | hCoV-19/England/ALDP-9C57FA/2020\|EPI_ISL_567535\|20 | ···················· | ···················· | ···················· | ···················· | ···················· | ···················· |
| K | 5033 | hCoV-19/England/ALDP-9C597C/2020\|EPI_ISL_567537\|20 | ···················· | ···················· | ···················· | ···················· | ···················· | ···················· |
| K | 5034 | hCoV-19/England/ALDP-9C54B7/2020\|EPI_ISL_567538\|20 | ···················· | ···················· | ···················· | ···················· | ···················· | ···················· |
| K | 5035 | hCoV-19/England/ALDP-9C5D9E/2020\|EPI_ISL_567539\|20 | ···················· | ···················· | ···················· | ···················· | ···················· | ···················· |
| K | 5036 | hCoV-19/England/ALDP-9C4B77/2020\|EPI_ISL_567540\|20 | ···················· | ···················· | ···················· | ···················· | ···················· | ···················· |
| K | 5037 | hCoV-19/England/ALDP-9C5E6E/2020\|EPI_ISL_567541\|20 | ···················· | ·A·················· | ···················· | ···················· | ···················· | ···················· |
| K | 5038 | hCoV-19/England/ALDP-9C5E5F/2020\|EPI_ISL_567542\|20 | ···················· | ·A·················· | ···················· | ···················· | ···················· | ···A················ |
| K | 5039 | hCoV-19/England/ALDP-9C55E1/2020\|EPI_ISL_567543\|20 | ···················· | ···················· | ···················· | ···················· | ···················· | ···················· |
| K | 5040 | hCoV-19/England/ALDP-9C60FE/2020\|EPI_ISL_567544\|20 | ···················· | ···················· | ···················· | ···················· | ···················· | ···················· |
| K | 5041 | hCoV-19/England/ALDP-9C51B0/2020\|EPI_ISL_567545\|20 | ···················· | ···················· | ···················· | ···················· | ···················· | ···················· |
| K | 5042 | hCoV-19/England/ALDP-9C59A9/2020\|EPI_ISL_567546\|20 | ···················· | ···················· | ···················· | ···················· | ···················· | ···················· |
| K | 5043 | hCoV-19/England/ALDP-9C5BFE/2020\|EPI_ISL_567547\|20 | ···················· | ···················· | ···················· | ···················· | ···················· | ···················· |
| K | 5044 | hCoV-19/England/ALDP-9C4C29/2020\|EPI_ISL_567548\|20 | ···················· | ···················· | ···················· | ···················· | ···················· | ···················· |
| K | 5045 | hCoV-19/England/ALDP-9C52DB/2020\|EPI_ISL_567549\|20 | ···················· | ···················· | ···················· | ···················· | ···················· | ···················· |
| K | 5046 | hCoV-19/England/ALDP-9C52F9/2020\|EPI_ISL_567550\|20 | ···················· | ···················· | ···················· | ···················· | ···················· | ···················· |
| K | 5047 | hCoV-19/England/ALDP-9C5648/2020\|EPI_ISL_567551\|20 | ···················· | ···················· | ···················· | ···················· | ······T············· | ···················· |
| K | 5048 | hCoV-19/England/ALDP-9C5B1C/2020\|EPI_ISL_567552\|20 | ···················· | ···················· | ···················· | ···················· | ···················· | ···················· |
| K | 5049 | hCoV-19/England/ALDP-9C560C/2020\|EPI_ISL_567553\|20 | ···················· | ···················· | ···················· | ···················· | ···················· | ···················· |
| K | 5050 | hCoV-19/England/ALDP-9C4EAB/2020\|EPI_ISL_567554\|20 | ···················· | ···················· | ···················· | ···················· | ···················· | ···················· |
| K | 5051 | hCoV-19/England/ALDP-9C637D/2020\|EPI_ISL_567555\|20 | ···················· | ···················· | ···················· | ···················· | ···················· | ···················· |
| K | 5052 | hCoV-19/England/ALDP-9C561B/2020\|EPI_ISL_567556\|20 | ···················· | ···················· | ···················· | ···················· | ···················· | ···················· |
| K | 5053 | hCoV-19/England/ALDP-9C4E23/2020\|EPI_ISL_567557\|20 | ···················· | ···················· | ···················· | ···················· | ···················· | ···················· |
| K | 5054 | hCoV-19/England/ALDP-9C50C2/2020\|EPI_ISL_567558\|20 | ···················· | ···················· | ···················· | ···················· | ····T··············· | ···················· |
| K | 5055 | hCoV-19/England/ALDP-9C56C0/2020\|EPI_ISL_567559\|20 | ···················· | ···················· | ···················· | ···················· | ···················· | ···················· |
| K | 5056 | hCoV-19/England/ALDP-9C5C19/2020\|EPI_ISL_567561\|20 | ···················· | ·A·················· | ···················· | ···················· | ···················· | ···················· |
| K | 5057 | hCoV-19/England/ALDP-9C56A2/2020\|EPI_ISL_567563\|20 | ···················· | ···················· | ···················· | ···················· | ···················· | ···················· |
| K | 5058 | hCoV-19/England/ALDP-9C4E7E/2020\|EPI_ISL_567564\|20 | ···················· | ···················· | ···················· | ···················· | ···················· | ···················· |
| K | 5059 | hCoV-19/England/ALDP-9C5E8C/2020\|EPI_ISL_567565\|20 | ···················· | ···················· | ···················· | ···················· | ···················· | ···················· |
| K | 5060 | hCoV-19/England/ALDP-9C5675/2020\|EPI_ISL_567566\|20 | ···················· | ···················· | ···················· | ···················· | ···················· | ···················· |
| K | 5061 | hCoV-19/England/ALDP-9C5A88/2020\|EPI_ISL_567567\|20 | ···················· | ···················· | ···················· | ···················· | ···················· | ···················· |
| K | 5062 | hCoV-19/England/ALDP-9C5921/2020\|EPI_ISL_567568\|20 | ········T··········· | ···················· | ···················· | ···················· | ···················· | ···················· |
| K | 5063 | hCoV-19/Wales/ALDP-9C5156/2020\|EPI_ISL_567569\|2020 | ···················· | ···················· | ···················· | ···················· | ···················· | ···················· |
| K | 5064 | hCoV-19/England/ALDP-9C5AD3/2020\|EPI_ISL_567570\|20 | ···················· | ···················· | ···················· | ···················· | ···················· | ···················· |
| K | 5065 | hCoV-19/England/ALDP-9C63AA/2020\|EPI_ISL_567571\|20 | ···················· | ···················· | ···················· | ···················· | ···················· | ···················· |
| K | 5066 | hCoV-19/England/ALDP-9C538D/2020\|EPI_ISL_567572\|20 | ···················· | ···················· | ···················· | ···················· | ···················· | ···················· |
| K | 5067 | hCoV-19/England/ALDP-9C5BC1/2020\|EPI_ISL_567573\|20 | ···················· | ···················· | ···················· | ···················· | ···················· | ···················· |
| K | 5068 | hCoV-19/England/ALDP-9C63C8/2020\|EPI_ISL_567574\|20 | ···················· | ···················· | ···················· | ···················· | ···················· | ···················· |
| K | 5069 | hCoV-19/England/ALDP-9C562A/2020\|EPI_ISL_567575\|20 | ···················· | ·A·················· | ···················· | ···················· | ···················· | ···A················ |
| K | 5070 | hCoV-19/England/ALDP-9C5341/2020\|EPI_ISL_567576\|20 | ···················· | ···················· | ···················· | ···················· | ···················· | ···················· |
| K | 5071 | hCoV-19/England/ALDP-9C61DD/2020\|EPI_ISL_567577\|20 | ···················· | ···················· | ···················· | ···················· | ···················· | ···················· |
| K | 5072 | hCoV-19/England/ALDP-9C4EBA/2020\|EPI_ISL_567578\|20 | ···················· | ···················· | ···················· | ···················· | ···················· | ···················· |
| K | 5073 | hCoV-19/England/ALDP-9C5842/2020\|EPI_ISL_567579\|20 | ···················· | ···················· | ···················· | ···················· | ···················· | ···················· |
| K | 5074 | hCoV-19/England/ALDP-9C5754/2020\|EPI_ISL_567722\|20 | ···················· | ···················· | ···················· | ···················· | ···················· | ···················· |
| K | 5075 | hCoV-19/England/ALDP-9C5745/2020\|EPI_ISL_567734\|20 | ···················· | ·A·················· | ···················· | ···················· | ···················· | ···A················ |
| K | 5076 | hCoV-19/England/ALDP-9C6067/2020\|EPI_ISL_567735\|20 | ···················· | ···················· | ···················· | ···················· | ···················· | ···················· |
| K | 5077 | hCoV-19/England/ALDP-9C5806/2020\|EPI_ISL_567736\|20 | ···················· | ···················· | ···················· | ···················· | ······T············· | ···················· |
| K | 5078 | hCoV-19/England/ALDP-9C4B95/2020\|EPI_ISL_567737\|20 | ···················· | ···················· | ···················· | ···················· | ···················· | ···················· |
| K | 5079 | hCoV-19/England/ALDP-9C587F/2020\|EPI_ISL_567739\|20 | ···················· | ···················· | ···················· | ···················· | ···················· | ···················· |
| K | 5080 | hCoV-19/England/ALDP-9C59C7/2020\|EPI_ISL_567740\|20 | ···················· | ···················· | ···················· | ···················· | ···················· | ···················· |
| K | 5081 | hCoV-19/England/ALDP-9C543F/2020\|EPI_ISL_567741\|20 | ···················· | ···················· | ···················· | ···················· | ···················· | ···················· |
| K | 5082 | hCoV-19/England/ALDP-9C5E7D/2020\|EPI_ISL_567742\|20 | ···················· | ···················· | ···················· | ···················· | ···················· | ···················· |
| K | 5083 | hCoV-19/England/ALDP-9C56FD/2020\|EPI_ISL_567743\|20 | ···················· | ···················· | ···················· | ···················· | ···················· | ···················· |
| K | 5084 | hCoV-19/England/ALDP-9C5A2E/2020\|EPI_ISL_567744\|20 | ···················· | ···················· | ···················· | ···················· | ···················· | ···················· |

| | | | | | | | |
|---|---|---|---|---|---|---|---|
| K | 5085 hCoV-19/England/ALDP-9C5305/2020\|EPI_ISL_567745\|20 | ···················· | ·A·················· | ···················· | ···················· | ···················· | ···A················ |
| K | 5086 hCoV-19/England/ALDP-9C553C/2020\|EPI_ISL_567746\|20 | ···················· | ···················· | ···················· | ···················· | ···················· | ···················· |
| K | 5087 hCoV-19/England/ALDP-9C4DF9/2020\|EPI_ISL_567747\|20 | ···················· | ···················· | ···················· | ···················· | ···················· | ···················· |
| K | 5088 hCoV-19/England/ALDP-9C601C/2020\|EPI_ISL_567748\|20 | ···················· | ·A·················· | ···················· | ···················· | ···················· | ···A················ |
| K | 5089 hCoV-19/England/ALDP-9C5AA6/2020\|EPI_ISL_567749\|20 | ···················· | ···················· | ···················· | ···················· | ···················· | ···················· |
| K | 5090 hCoV-19/England/ALDP-9C52EA/2020\|EPI_ISL_567750\|20 | ···················· | ···················· | ···················· | ···················· | ···················· | ···················· |
| K | 5091 hCoV-19/England/ALDP-9C5129/2020\|EPI_ISL_567751\|20 | ···················· | ···················· | ···················· | ···················· | ···················· | ···················· |
| K | 5092 hCoV-19/England/ALDP-9C510B/2020\|EPI_ISL_567753\|20 | ···················· | ···················· | ···················· | ···················· | ······T············· | ···················· |
| K | 5093 hCoV-19/England/ALDP-9C6094/2020\|EPI_ISL_567754\|20 | ···················· | ·A·················· | ···················· | ···················· | ···················· | ···A················ |
| K | 5094 hCoV-19/England/ALDP-9C5AB5/2020\|EPI_ISL_567755\|20 | ···················· | ···················· | ···················· | ···················· | ···················· | ···················· |
| K | 5095 hCoV-19/Kenya/C7605/2020\|EPI_ISL_568847\|2020-05-16 | ···················· | ···················· | ···················· | ···················· | ···················· | ···················· |
| K | 5096 hCoV-19/Kenya/C21582/2020\|EPI_ISL_568872\|2020-06-2 | ···················· | ···················· | ···················· | ···················· | ···················· | ···················· |
| K | 5097 hCoV-19/Malaysia/MGI-M23/2020\|EPI_ISL_568873\|2020- | ···················· | ···················· | ···················· | ···················· | ···················· | ···················· |
| K | 5098 hCoV-19/Malaysia/MGI-G812/2020\|EPI_ISL_568874\|2020 | ···················· | ···················· | ···················· | ···················· | ···················· | ···················· |
| K | 5099 hCoV-19/Malaysia/MGI-MAEPS126/2020\|EPI_ISL_568875\| | ···················· | ···················· | ···················· | ···················· | ···················· | ···················· |
| K | 5100 hCoV-19/France/PAC-IHU-0593/2020\|EPI_ISL_568877\|20 | ···················· | ···················· | ···················· | ···················· | ···················· | ···················· |
| K | 5101 hCoV-19/France/PAC-IHU-0594/2020\|EPI_ISL_568878\|20 | ···················· | ···················· | ···················· | ···················· | ···················· | ···················· |
| K | 5102 hCoV-19/France/PAC-IHU-0595/2020\|EPI_ISL_568879\|20 | ···················· | ···················· | ···················· | ···················· | ···················· | ···················· |
| K | 5103 hCoV-19/France/PAC-IHU-0597/2020\|EPI_ISL_568880\|20 | ···················· | ···················· | ···················· | ···················· | ···················· | ···················· |
| K | 5104 hCoV-19/France/PAC-IHU-0601/2020\|EPI_ISL_568881\|20 | ···················· | ···················· | ···················· | ···················· | ···················· | ···················· |
| K | 5105 hCoV-19/France/PAC-IHU-0602/2020\|EPI_ISL_568882\|20 | ···················· | ···················· | ···················· | ···················· | ···················· | ···················· |
| K | 5106 hCoV-19/France/PAC-IHU-0603/2020\|EPI_ISL_568883\|20 | ···················· | ···················· | ···················· | ···················· | ···················· | ···················· |
| K | 5107 hCoV-19/France/PAC-IHU-0604/2020\|EPI_ISL_568884\|20 | ···················· | ···················· | ···················· | ···················· | ···················· | ···················· |
| K | 5108 hCoV-19/France/PAC-IHU-0609/2020\|EPI_ISL_568885\|20 | ···················· | ···················· | ···················· | ···················· | ···················· | ···················· |
| K | 5109 hCoV-19/France/PAC-IHU-0610/2020\|EPI_ISL_568886\|20 | ···················· | ···················· | ···················· | ···················· | ···················· | ···················· |
| K | 5110 hCoV-19/France/PAC-IHU-0611/2020\|EPI_ISL_568887\|20 | ···················· | ···················· | ···················· | ···················· | ···················· | ···················· |
| K | 5111 hCoV-19/France/PAC-IHU-0612/2020\|EPI_ISL_568888\|20 | ···················· | ···················· | ···················· | ···················· | ···················· | ···················· |
| K | 5112 hCoV-19/France/PAC-IHU-0615/2020\|EPI_ISL_568889\|20 | ···················· | ···················· | ···················· | ···················· | ···················· | ···················· |
| K | 5113 hCoV-19/France/PAC-IHU-0619/2020\|EPI_ISL_568892\|20 | ···················· | ···················· | ···················· | ···················· | ···················· | ···················· |
| K | 5114 hCoV-19/France/PAC-IHU-0620/2020\|EPI_ISL_568893\|20 | ···················· | ···················· | ···················· | ···················· | ···················· | ···················· |
| K | 5115 hCoV-19/France/PAC-IHU-0621/2020\|EPI_ISL_568894\|20 | ···················· | ···················· | ···················· | ···················· | ···················· | ···················· |
| K | 5116 hCoV-19/France/PAC-IHU-0622/2020\|EPI_ISL_568895\|20 | ···················· | ···················· | ···················· | ···················· | ···················· | ···················· |
| K | 5117 hCoV-19/France/PAC-IHU-0623/2020\|EPI_ISL_568896\|20 | ···················· | ···················· | ···················· | ···················· | ···················· | ···················· |
| K | 5118 hCoV-19/France/PAC-IHU-0624/2020\|EPI_ISL_568897\|20 | ···················· | ···················· | ···················· | ···················· | ···················· | ···················· |
| K | 5119 hCoV-19/France/PAC-IHU-0627/2020\|EPI_ISL_568898\|20 | ···················· | ···················· | ···················· | ···················· | ···················· | ···················· |
| K | 5120 hCoV-19/France/PAC-IHU-0628/2020\|EPI_ISL_568899\|20 | ···················· | ···················· | ···················· | ···················· | ···················· | ···················· |
| K | 5123 hCoV-19/Australia/VIC9510/2020\|EPI_ISL_562742\|2020 | ···················· | ···················· | ···················· | ···················· | ···················· | ···················· |
| K | 5124 hCoV-19/Australia/VIC9532/2020\|EPI_ISL_562743\|2020 | ···················· | ···················· | ···················· | ···················· | ···················· | ···················· |
| K | 5125 hCoV-19/Australia/VIC9633/2020\|EPI_ISL_562744\|2020 | ···················· | ···················· | ···················· | ···················· | ···················· | ···················· |
| K | 5126 hCoV-19/Australia/VIC9634/2020\|EPI_ISL_562745\|2020 | ···················· | ···················· | ···················· | ···················· | ···················· | ···················· |
| K | 5127 hCoV-19/Australia/VIC9635/2020\|EPI_ISL_562746\|2020 | ···················· | ···················· | ···················· | ···················· | ···················· | ···················· |
| K | 5128 hCoV-19/Australia/VIC9636/2020\|EPI_ISL_562747\|2020 | ···················· | ···················· | ···················· | ···················· | ···················· | ···················· |
| K | 5129 hCoV-19/Australia/VIC9637/2020\|EPI_ISL_562748\|2020 | ···················· | ···················· | ···················· | ···················· | ···················· | ···················· |
| K | 5130 hCoV-19/Australia/VIC9884/2020\|EPI_ISL_562749\|2020 | ···················· | ···················· | ···················· | ···················· | ···················· | ···················· |
| K | 5132 hCoV-19/Australia/VIC9987/2020\|EPI_ISL_562751\|2020 | ···················· | ···················· | ···················· | ···················· | ···················· | ···················· |
| K | 5133 hCoV-19/Australia/VIC9988/2020\|EPI_ISL_562752\|2020 | ···················· | ···················· | ···················· | ···················· | ···················· | ···················· |
| K | 5134 hCoV-19/Australia/VIC9990/2020\|EPI_ISL_562753\|2020 | ···················· | ···················· | ···················· | ···················· | ···················· | ···················· |
| K | 5135 hCoV-19/Australia/VIC10004/2020\|EPI_ISL_562761\|202 | ···················· | ···················· | ···················· | ···················· | ···················· | ···················· |
| K | 5136 hCoV-19/Australia/VIC10006/2020\|EPI_ISL_562762\|202 | ···················· | ···················· | ···················· | ···················· | ···················· | ···················· |
| K | 5137 hCoV-19/Australia/VIC10017/2020\|EPI_ISL_562763\|202 | ···················· | ···················· | ···················· | ···················· | ···················· | ···················· |
| K | 5138 hCoV-19/Australia/VIC10019/2020\|EPI_ISL_562764\|202 | ···················· | ···················· | ···················· | ···················· | ···················· | ···················· |
| K | 5139 hCoV-19/Australia/VIC10020/2020\|EPI_ISL_562765\|202 | ···················· | ···················· | ···················· | ···················· | ···················· | ···················· |
| K | 5140 hCoV-19/Australia/VIC10024/2020\|EPI_ISL_562766\|202 | ···················· | ···················· | ···················· | ···················· | ···················· | ···················· |
| K | 5141 hCoV-19/Australia/VIC10025/2020\|EPI_ISL_562767\|202 | ···················· | ···················· | ···················· | ···················· | ···················· | ···················· |
| K | 5142 hCoV-19/Australia/VIC10026/2020\|EPI_ISL_562768\|202 | ···················· | ···················· | ···················· | ···················· | ···················· | ···················· |
| K | 5143 hCoV-19/Australia/VIC10029/2020\|EPI_ISL_562769\|202 | ···················· | ···················· | ···················· | ···················· | ···················· | ···················· |
| K | 5144 hCoV-19/Australia/VIC10031/2020\|EPI_ISL_562770\|202 | ···················· | ···················· | ···················· | ···················· | ···················· | ···················· |
| K | 5145 hCoV-19/Australia/VIC10032/2020\|EPI_ISL_562771\|202 | ···················· | ···················· | ···················· | ···················· | ···················· | ···················· |
| K | 5146 hCoV-19/Australia/VIC10035/2020\|EPI_ISL_562772\|202 | ···················· | ···················· | ···················· | ···················· | ···················· | ···················· |
| K | 5147 hCoV-19/Australia/VIC10037/2020\|EPI_ISL_562773\|202 | ···················· | ···················· | ···················· | ···················· | ···················· | ···················· |
| K | 5148 hCoV-19/Australia/VIC10041/2020\|EPI_ISL_562774\|202 | ···················· | ···················· | ···················· | ···················· | ···················· | ···················· |
| K | 5149 hCoV-19/Australia/VIC10043/2020\|EPI_ISL_562775\|202 | ···················· | ···················· | ···················· | ···················· | ···················· | ···················· |
| K | 5150 hCoV-19/Australia/VIC10045/2020\|EPI_ISL_562776\|202 | ···················· | ···················· | ···················· | ···················· | ···················· | ···················· |
| K | 5151 hCoV-19/Australia/VIC10051/2020\|EPI_ISL_562777\|202 | ···················· | ···················· | ···················· | ···················· | ···················· | ···················· |
| K | 5152 hCoV-19/Australia/VIC10055/2020\|EPI_ISL_562778\|202 | ···················· | ···················· | ···················· | ···················· | ···················· | ···················· |
| K | 5153 hCoV-19/Australia/VIC10060/2020\|EPI_ISL_562779\|202 | ···················· | ···················· | ···················· | ···················· | ···················· | ···················· |
| K | 5154 hCoV-19/Australia/VIC10061/2020\|EPI_ISL_562780\|202 | ···················· | ···················· | ···················· | ···················· | ···················· | ···················· |
| K | 5155 hCoV-19/Australia/VIC10065/2020\|EPI_ISL_562781\|202 | ···················· | ···················· | ···················· | ···················· | ···················· | ···················· |
| K | 5156 hCoV-19/Australia/VIC10066/2020\|EPI_ISL_562782\|202 | ···················· | ···················· | ···················· | ···················· | ···················· | ···················· |
| K | 5157 hCoV-19/Australia/VIC10067/2020\|EPI_ISL_562783\|202 | ···················· | ···················· | ···················· | ···················· | ···················· | ···················· |
| K | 5158 hCoV-19/Australia/VIC10068/2020\|EPI_ISL_562784\|202 | ···················· | ···················· | ···················· | ···················· | ···················· | ···················· |
| K | 5159 hCoV-19/Australia/VIC10069/2020\|EPI_ISL_562785\|202 | ···················· | ···················· | ···················· | ···················· | ···················· | ···················· |
| K | 5160 hCoV-19/Australia/VIC10075/2020\|EPI_ISL_562786\|202 | ···················· | ···················· | ···················· | ···················· | ···················· | ···················· |
| K | 5161 hCoV-19/Australia/VIC10080/2020\|EPI_ISL_562787\|202 | ···················· | ···················· | ···················· | ···················· | ···················· | ···················· |
| K | 5162 hCoV-19/Australia/VIC10081/2020\|EPI_ISL_562788\|202 | ···················· | ···················· | ···················· | ···················· | ···················· | ···················· |
| K | 5163 hCoV-19/Australia/VIC10085/2020\|EPI_ISL_562789\|202 | ···················· | ···················· | ···················· | ···················· | ···················· | ···················· |
| K | 5164 hCoV-19/Australia/VIC10093/2020\|EPI_ISL_562790\|202 | ···················· | ···················· | ···················· | ···················· | ···················· | ···················· |
| K | 5165 hCoV-19/Australia/VIC10095/2020\|EPI_ISL_562791\|202 | ···················· | ···················· | ···K················ | ···················· | ···················· | ···················· |
| K | 5166 hCoV-19/Australia/VIC10098/2020\|EPI_ISL_562792\|202 | ···················· | ···················· | ···N················ | ···················· | ···················· | ·····N·············· |
| K | 5167 hCoV-19/Australia/VIC10115/2020\|EPI_ISL_562793\|202 | ···················· | ···················· | ···················· | ···················· | ···················· | ···················· |

K 5168 hCoV-19/Australia/VIC10116/2020|EPI_ISL_562794|202 ···················· ···················· ····················· ···················· ···················· ····················

K 5169 hCoV-19/Australia/VIC10118/2020|EPI_ISL_562795|202 ···················· ···················· ····················· ···················· ···················· ····················

K 5170 hCoV-19/Australia/VIC10120/2020|EPI_ISL_562796|202 ···················· ···················· ····················· ···················· ···················· ····················

K 5171 hCoV-19/Australia/VIC10121/2020|EPI_ISL_562797|202 ···················· ···················· ····················· ···················· ···················· ····················

K 5172 hCoV-19/Australia/VIC10122/2020|EPI_ISL_562798|202 ···················· ···················· ····················· ···················· ···················· ····················

K 5173 hCoV-19/Australia/VIC10125/2020|EPI_ISL_562799|202 ···················· ···················· ····················· ···················· ···················· ····················

K 5174 hCoV-19/Australia/VIC13967/2020|EPI_ISL_563900|202 ···················· ···················· ····················· ···················· ···················· ····················

K 5175 hCoV-19/Australia/VIC13968/2020|EPI_ISL_563901|202 ···················· ···················· ····················· ···················· ···················· ····················

K 5177 hCoV-19/Australia/VIC13971/2020|EPI_ISL_563903|202 ···················· ···················· ····················· ···················· ···················· ····················

K 5178 hCoV-19/Australia/VIC13975/2020|EPI_ISL_563904|202 ···················· ···················· ····················· ···················· ···················· ····················

K 5183 hCoV-19/Australia/VIC13984/2020|EPI_ISL_563909|202 ···················· ···················· ····················· ···················· ···················· ····················

K 5184 hCoV-19/Australia/VIC14004/2020|EPI_ISL_563921|202 ···················· ···················· ····················· ···················· ···················· ····················

K 5186 hCoV-19/Australia/VIC14008/2020|EPI_ISL_563923|202 ···················· ···················· ····················· ···················· ···················· ····················

K 5188 hCoV-19/Australia/VIC14010/2020|EPI_ISL_563926|202 ···················· ···················· ····················· ···················· ···················· ····················

K 5189 hCoV-19/Australia/VIC14011/2020|EPI_ISL_563927|202 ···················· ···················· ····················· ···················· ···················· ····················

K 5190 hCoV-19/Australia/VIC14013/2020|EPI_ISL_563928|202 ···················· ···················· ····················· ···················· ···················· ····················

K 5191 hCoV-19/Australia/VIC14014/2020|EPI_ISL_563929|202 ···················· ···················· ····················· ···················· ···················· ····················

K 5192 hCoV-19/England/ALDP-9CD6AA/2020|EPI_ISL_566151|20 ···················· ···················· ····················· ···················· ···················· ····················

K 5193 hCoV-19/England/ALDP-9CD58F/2020|EPI_ISL_566153|20 ···················· ·A·················· ····················· ···················· ···················· ···A················

K 5194 hCoV-19/England/ALDP-9CD40A/2020|EPI_ISL_566154|20 ···················· ···················· ····················· ···················· ···················· ····················

K 5195 hCoV-19/England/ALDP-9CDCA8/2020|EPI_ISL_566155|20 ···················· ···················· ····················· ···················· ···················· ····················

K 5196 hCoV-19/England/ALDP-9CD69B/2020|EPI_ISL_566156|20 ···················· ···················· ····················· ···················· ···················· ····················

K 5197 hCoV-19/England/ALDP-9CD5BC/2020|EPI_ISL_566157|20 ···················· ···················· ····················· ···················· ···················· ····················

K 5198 hCoV-19/England/ALDP-9CDA26/2020|EPI_ISL_566159|20 ···················· ···················· ····················· ···················· ···················· ····················

K 5199 hCoV-19/England/ALDP-9CD6D7/2020|EPI_ISL_566160|20 ···················· ···················· ····················· ···················· ···················· ····················

K 5200 hCoV-19/England/ALDP-9CD24C/2020|EPI_ISL_566161|20 ···················· ···················· ····················· ···················· ···················· ····················

K 5201 hCoV-19/England/ALDP-9CD5E9/2020|EPI_ISL_566162|20 ···················· ·A·················· ····················· ···················· ···················· ···A················

K 5202 hCoV-19/England/ALDP-9CD77A/2020|EPI_ISL_566163|20 ···················· ···················· ····················· ···················· ···················· ····················

K 5203 hCoV-19/England/ALDP-9CD7A7/2020|EPI_ISL_566164|20 ···················· ···················· ····················· ···················· ···················· ····················

K 5204 hCoV-19/England/ALDP-9CD16D/2020|EPI_ISL_566166|20 ···················· ···················· ····················· ···················· ····T··············· ····················

K 5205 hCoV-19/England/ALDP-9CD525/2020|EPI_ISL_566167|20 ···················· ·R·················· ····················· ···················· ···················· ···R················

K 5206 hCoV-19/England/ALDP-9CDD5A/2020|EPI_ISL_566168|20 ···················· ···················· ····················· ···················· ···················· ····················

K 5207 hCoV-19/England/ALDP-9CCDE2/2020|EPI_ISL_566169|20 ···················· ···················· ····················· ···················· ···················· ····················

K 5208 hCoV-19/England/ALDP-9CD68C/2020|EPI_ISL_566170|20 ···················· ···················· ····················· ···················· ···················· ····················

K 5209 hCoV-19/England/ALDP-9CDD78/2020|EPI_ISL_566171|20 ···················· ···················· ····················· ···················· ···················· ····················

K 5210 hCoV-19/England/ALDP-9CD534/2020|EPI_ISL_566174|20 ···················· ···················· ····················· ···················· ···················· ····················

K 5211 hCoV-19/England/ALDP-9CDB05/2020|EPI_ISL_566175|20 ···················· ···················· ····················· ···················· ···················· ····················

K 5212 hCoV-19/England/ALDP-9CD279/2020|EPI_ISL_566176|20 ···················· ···················· ····················· ···················· ···················· ····················

K 5213 hCoV-19/England/ALDP-9CDC02/2020|EPI_ISL_566177|20 ···················· ···················· ····················· ···················· ···················· ····················

K 5214 hCoV-19/England/ALDP-9CD9ED/2020|EPI_ISL_566179|20 ···················· ···················· ····················· ···················· ···················· ····················

K 5215 hCoV-19/England/ALDP-9CD8C2/2020|EPI_ISL_566180|20 ···················· ···················· ····················· ···················· ···················· ····················

K 5216 hCoV-19/England/ALDP-9CD060/2020|EPI_ISL_566181|20 ···················· ···················· ····················· ···················· ···················· ····················

K 5217 hCoV-19/England/ALDP-9CCE0D/2020|EPI_ISL_566182|20 ···················· ···················· ····················· ···················· ···················· ····················

K 5218 hCoV-19/England/ALDP-9CE199/2020|EPI_ISL_566183|20 ···················· ···················· ····················· ···················· ···················· ····················

K 5219 hCoV-19/England/ALDP-9CD2D3/2020|EPI_ISL_566184|20 ···················· ·A·················· ····················· ···················· ···················· ···A················

K 5220 hCoV-19/England/ALDP-9CDA71/2020|EPI_ISL_566185|20 ···················· ·A·················· ····················· ···················· ···················· ···A················

K 5221 hCoV-19/England/ALDP-9CD631/2020|EPI_ISL_566186|20 ···················· ·A·················· ····················· ···················· ···················· ···A················

K 5222 hCoV-19/England/ALDP-9CD710/2020|EPI_ISL_566187|20 ···················· ···················· ····················· ···················· ···················· ····················

K 5223 hCoV-19/England/ALDP-9CD5AD/2020|EPI_ISL_566188|20 ···················· ···················· ····················· ···················· ···················· ····················

K 5224 hCoV-19/England/ALDP-9CD33A/2020|EPI_ISL_566189|20 ···················· ···················· ····················· ···················· ···················· ····················

K 5225 hCoV-19/England/ALDP-9CCD2E/2020|EPI_ISL_566249|20 ···················· ·A·················· ····················· ···················· ···················· ···A················

K 5226 hCoV-19/England/ALDP-9CD640/2020|EPI_ISL_566250|20 ···················· ···················· ····················· ···················· ···················· ····················

K 5227 hCoV-19/England/ALDP-9CD9A1/2020|EPI_ISL_566251|20 ···················· ···················· ····················· ···················· ···················· ····················

K 5228 hCoV-19/England/ALDP-9CDAF9/2020|EPI_ISL_566252|20 ···················· ···················· ····················· ···················· ···················· ····················

K 5229 hCoV-19/England/ALDP-9CCDF1/2020|EPI_ISL_566253|20 ···················· ···················· ····················· ···················· ···················· ····················

K 5230 hCoV-19/England/ALDP-9CD385/2020|EPI_ISL_566255|20 ···················· ···················· ····················· ···················· ···················· ····················

K 5231 hCoV-19/England/ALDP-9CDD0F/2020|EPI_ISL_566256|20 ···················· ···················· ····················· ···················· ···················· ····················

K 5232 hCoV-19/England/ALDP-9CD1C7/2020|EPI_ISL_566257|20 ···················· ···················· ····················· ···················· ···················· ····················

K 5233 hCoV-19/England/ALDP-9CD4A0/2020|EPI_ISL_566258|20 ···················· ···················· ····················· ···················· ···················· ····················

K 5234 hCoV-19/England/ALDP-9CD868/2020|EPI_ISL_566259|20 ···················· ···················· ····················· ···················· ···················· ····················

K 5235 hCoV-19/England/ALDP-9CCE85/2020|EPI_ISL_566260|20 ···················· ···················· ····················· ···················· ···················· ····················

K 5236 hCoV-19/England/ALDP-9CD561/2020|EPI_ISL_566261|20 ···················· ···················· ····················· ···················· ···················· ····················

K 5237 hCoV-19/England/ALDP-9CD428/2020|EPI_ISL_566262|20 ···················· ···················· ····················· ···················· ···················· ····················

K 5238 hCoV-19/England/ALDP-9CD0E8/2020|EPI_ISL_566263|20 ···················· ···················· ····················· ···················· ···················· ····················

K 5239 hCoV-19/England/ALDP-9CDBC9/2020|EPI_ISL_566264|20 ···················· ···················· ····················· ···················· ···················· ····················

K 5240 hCoV-19/England/ALDP-9CDBE7/2020|EPI_ISL_566265|20 ···················· ···················· ····················· ···················· ···················· ····················

K 5241 hCoV-19/England/ALDP-9CDAAE/2020|EPI_ISL_566267|20 ···················· ···················· ····················· ···················· ···················· ····················

K 5242 hCoV-19/England/ALDP-9CD938/2020|EPI_ISL_566268|20 ···················· ···················· ····················· ···················· ···················· ····················

K 5243 hCoV-19/England/ALDP-9CD0D9/2020|EPI_ISL_566269|20 ···················· ···················· ····················· ···················· ···················· ····················

K 5244 hCoV-19/England/ALDP-9CD895/2020|EPI_ISL_566300|20 ···················· ···················· ····················· ···················· ···················· ····················

K 5245 hCoV-19/England/ALDP-9CD72F/2020|EPI_ISL_566301|20 ···················· ···················· ·····G··············· ···················· ···················· ····················

K 5246 hCoV-19/England/ALDP-9CDD4B/2020|EPI_ISL_566302|20 ···················· ···················· ····················· ···················· ···················· ····················

K 5247 hCoV-19/England/ALDP-9CD965/2020|EPI_ISL_566303|20 ···················· ···················· ····················· ···················· ···················· ····················

K 5248 hCoV-19/England/QEUH-9CA7D7/2020|EPI_ISL_567480|20 ···················· ···················· ····················· ···················· ···················· ····················

K 5249 hCoV-19/England/QEUH-9C9D4F/2020|EPI_ISL_567481|20 ···················· ·A·················· ····················· ···················· ···················· ···A················

K 5250 hCoV-19/England/QEUH-9CA40D/2020|EPI_ISL_567482|20 ···················· ···················· ····················· ···················· ···················· ····················

K 5251 hCoV-19/England/QEUH-9C9574/2020|EPI_ISL_567483|20 ···················· ···················· ····················· ···················· ···················· ····················

K 5252 hCoV-19/England/QEUH-9CA573/2020|EPI_ISL_567484|20 ···················· ···················· ····················· ···················· ···················· ····················

K 5253 hCoV-19/England/QEUH-9CA713/2020|EPI_ISL_567485|20 ···················· ···················· ····················· ···················· ···················· ····················

K 5254 hCoV-19/England/QEUH-9C9E1F/2020|EPI_ISL_567486|20 ···················· ···················· ····················· ···················· ···················· ····················

K 5255 hCoV-19/England/QEUH-9CA5CE/2020|EPI_ISL_567488|20 ···················· ·A·················· ···················· ···················· ···················· ····················
K 5256 hCoV-19/England/QEUH-9C991E/2020|EPI_ISL_567489|20 ···················· ···················· ···················· ···················· ···················· ····················
K 5257 hCoV-19/England/QEUH-9C9969/2020|EPI_ISL_567490|20 ···················· ···················· ···················· ···················· ···················· ····················
K 5258 hCoV-19/Wales/QEUH-9C9B90/2020|EPI_ISL_567492|2020 ···················· ···················· ···················· ···················· ···················· ····················
K 5259 hCoV-19/England/QEUH-9C98B7/2020|EPI_ISL_567493|20 ···················· ···················· ···················· ···················· ···················· ····················
K 5260 hCoV-19/England/QEUH-9CA43A/2020|EPI_ISL_567494|20 ···················· ···················· ···················· ···················· ···················· ····················
K 5261 hCoV-19/England/QEUH-9C9BAF/2020|EPI_ISL_567495|20 ···················· ···················· ···················· ···················· ····T··············· ····················
K 5262 hCoV-19/England/QEUH-9CA50A/2020|EPI_ISL_567496|20 ···················· ···················· ···················· ···················· ···················· ····················
K 5263 hCoV-19/England/QEUH-9C95A1/2020|EPI_ISL_567497|20 ···················· ·A·················· ···················· ···················· ···················· ····················
K 5264 hCoV-19/England/QEUH-9CA801/2020|EPI_ISL_567498|20 ···················· ···················· ···················· ···················· ···················· ····················
K 5265 hCoV-19/England/QEUH-9CA0EB/2020|EPI_ISL_567499|20 ···················· ···················· ···················· ···················· ···················· ····················
K 5266 hCoV-19/England/ALDP-9C5F2F/2020|EPI_ISL_567560|20 ···················· ···················· ···················· ···················· ···················· ····················
K 5267 hCoV-19/England/ALDP-9C5BA3/2020|EPI_ISL_567562|20 ···················· ···················· ···················· ···················· ···················· ····················
K 5268 hCoV-19/England/ALDP-9C4D44/2020|EPI_ISL_567580|20 ···················· ···················· ···················· ···················· ···················· ····················
K 5269 hCoV-19/England/ALDP-9C5F01/2020|EPI_ISL_567581|20 ···················· ···················· ···················· ···················· ···················· ····················
K 5270 hCoV-19/England/ALDP-9C5DDA/2020|EPI_ISL_567582|20 ···················· ···················· ···················· ···················· ···················· ····················
K 5271 hCoV-19/England/ALDP-9C51A1/2020|EPI_ISL_567583|20 ···················· ···················· ···················· ···················· ···················· ····················
K 5272 hCoV-19/England/ALDP-9C5D52/2020|EPI_ISL_567584|20 ···················· ···················· ···················· ···················· ···················· ····················
K 5273 hCoV-19/England/ALDP-9C53F6/2020|EPI_ISL_567585|20 ···················· ···················· ···················· ···················· ···················· ····················
K 5274 hCoV-19/England/ALDP-9C54A8/2020|EPI_ISL_567586|20 ···················· ···················· ···················· ···················· ···················· ····················
K 5275 hCoV-19/England/ALDP-9C50D1/2020|EPI_ISL_567587|20 ···················· ···················· ···················· ···················· ····T··············· ····················
K 5276 hCoV-19/England/ALDP-9C4F4E/2020|EPI_ISL_567589|20 ···················· ···················· ···················· ···················· ···················· ····················
K 5277 hCoV-19/England/ALDP-9C500E/2020|EPI_ISL_567590|20 ···················· ···················· ···················· ···················· ···················· ····················
K 5278 hCoV-19/England/ALDP-9C639B/2020|EPI_ISL_567591|20 ···················· ···················· ···················· ···················· ···················· ····················
K 5279 hCoV-19/England/ALDP-9C53E7/2020|EPI_ISL_567592|20 ···················· ·A·················· ···················· ···················· ···················· ···A················
K 5280 hCoV-19/England/ALDP-9C5CCE/2020|EPI_ISL_567595|20 ···················· ···················· ···················· ···················· ···················· ····················
K 5281 hCoV-19/England/ALDP-9C5684/2020|EPI_ISL_567596|20 ···················· ···················· ···················· ···················· ···················· ····················
K 5282 hCoV-19/England/ALDP-9C5077/2020|EPI_ISL_567597|20 ···················· ···················· ···················· ···················· ···················· ····················
K 5283 hCoV-19/England/ALDP-9C4F5D/2020|EPI_ISL_567598|20 ···················· ·A·················· ···················· ···················· ···················· ···A················
K 5284 hCoV-19/England/ALDP-9C5C0A/2020|EPI_ISL_567599|20 ···················· ···················· ···················· ···················· ···················· ····················
K 5285 hCoV-19/England/ALDP-9C5781/2020|EPI_ISL_567600|20 ···················· ···················· ···················· ···················· ···················· ····················
K 5286 hCoV-19/England/ALDP-9C55D2/2020|EPI_ISL_567601|20 ···················· ···················· ···················· ···················· ···················· ····················
K 5287 hCoV-19/England/ALDP-9C53C9/2020|EPI_ISL_567602|20 ···················· ···················· ···················· ···················· ···················· ····················
K 5288 hCoV-19/England/ALDP-9C5E31/2020|EPI_ISL_567604|20 ···················· ···················· ···················· ···················· ···················· ····················
K 5289 hCoV-19/England/ALDP-9C5709/2020|EPI_ISL_567605|20 ···················· ···················· ···················· ···················· ···················· ····················
K 5290 hCoV-19/England/ALDP-9C5EF5/2020|EPI_ISL_567606|20 ···················· ···················· ···················· ···················· ···················· ····················
K 5291 hCoV-19/England/ALDP-9C4CDE/2020|EPI_ISL_567608|20 ···················· ···················· ···················· ···················· ···················· ····················
K 5292 hCoV-19/England/ALDP-9C5B2B/2020|EPI_ISL_567609|20 ···················· ···················· ···················· ···················· ···················· ····················
K 5293 hCoV-19/England/ALDP-9C4BD1/2020|EPI_ISL_567610|20 ···················· ···················· ···················· ···················· ···················· ····················
K 5294 hCoV-19/England/ALDP-9C5226/2020|EPI_ISL_567611|20 ···················· ···················· ···················· ···················· ···················· ····················
K 5295 hCoV-19/England/ALDP-9C5E22/2020|EPI_ISL_567612|20 ···················· ···················· ···················· ···················· ···················· ····················
K 5296 hCoV-19/England/ALDP-9C55C3/2020|EPI_ISL_567613|20 ···················· ···················· ···················· ···················· ···················· ····················
K 5297 hCoV-19/England/ALDP-9C4BC2/2020|EPI_ISL_567614|20 ···················· ···················· ···················· ···················· ···················· ····················
K 5298 hCoV-19/England/ALDP-9C5815/2020|EPI_ISL_567615|20 ···················· ···················· ···················· ···················· ······T············· ····················
K 5299 hCoV-19/England/ALDP-9C56EE/2020|EPI_ISL_567616|20 ···················· ···················· ···················· ···················· ···················· ····················
K 5300 hCoV-19/England/ALDP-9C4D62/2020|EPI_ISL_567617|20 ···················· ·A·················· ···················· ···················· ···················· ···A················
K 5301 hCoV-19/England/ALDP-9C5BEF/2020|EPI_ISL_567618|20 ···················· ·A·················· ···················· ···················· ···················· ···A················
K 5302 hCoV-19/England/ALDP-9C5086/2020|EPI_ISL_567619|20 ···················· ···················· ···················· ···················· ···················· ····················
K 5303 hCoV-19/England/ALDP-9C6191/2020|EPI_ISL_567620|20 ···················· ···················· ···················· ···················· ···················· ····················
K 5304 hCoV-19/England/ALDP-9C5B94/2020|EPI_ISL_567621|20 ···················· ···················· ···················· ···················· ···················· ····················
K 5305 hCoV-19/England/ALDP-9C536F/2020|EPI_ISL_567622|20 ···················· ···················· ···················· ···················· ···················· ····················
K 5306 hCoV-19/England/ALDP-9C4C65/2020|EPI_ISL_567624|20 ···················· ···················· ···················· ···················· ···················· ····················
K 5307 hCoV-19/England/ALDP-9C52CC/2020|EPI_ISL_567625|20 ···················· ···················· ···················· ···················· ···················· ····················
K 5308 hCoV-19/England/ALDP-9C6401/2020|EPI_ISL_567626|20 ···················· ···················· ···················· ···················· ···················· ····················
K 5309 hCoV-19/England/ALDP-9C5903/2020|EPI_ISL_567627|20 ···················· ···················· ···················· ···················· ···················· ····················
K 5310 hCoV-19/England/ALDP-9C4E8D/2020|EPI_ISL_567628|20 ···················· ···················· ···················· ···················· ···················· ····················
K 5311 hCoV-19/England/ALDP-9C4E50/2020|EPI_ISL_567629|20 ···················· ···················· ···················· ···················· ···················· ····················
K 5312 hCoV-19/England/ALDP-9C5D07/2020|EPI_ISL_567630|20 ···················· ···················· ···················· ···················· ···················· ····················
K 5313 hCoV-19/England/ALDP-9C5E13/2020|EPI_ISL_567631|20 ···················· ···················· ···················· ···················· ···················· ····················
K 5314 hCoV-19/England/ALDP-9C4F11/2020|EPI_ISL_567632|20 ···················· ·A·················· ···················· ···················· ···················· ···A················
K 5315 hCoV-19/England/ALDP-9C57BE/2020|EPI_ISL_567633|20 ···················· ···················· ···················· ···················· ···················· ····················
K 5316 hCoV-19/England/ALDP-9C4EE7/2020|EPI_ISL_567634|20 ···················· ···················· ···················· ···················· ···················· ····················
K 5317 hCoV-19/France/PAC-IHU-0617/2020|EPI_ISL_568890|20 ···················· ···················· ···················· ···················· ···················· ····················
K 5318 hCoV-19/France/PAC-IHU-0618/2020|EPI_ISL_568891|20 ···················· ···················· ···················· ···················· ···················· ····················
K 5319 hCoV-19/France/PAC-IHU-0630/2020|EPI_ISL_568901|20 ···················· ···················· ···················· ···················· ···················· ····················
K 5320 hCoV-19/France/PAC-IHU-0631/2020|EPI_ISL_568902|20 ···················· ···················· ···················· ···················· ···················· ····················
K 5321 hCoV-19/France/PAC-IHU-0632/2020|EPI_ISL_568903|20 ···················· ···················· ···················· ···················· ···················· ····················
K 5322 hCoV-19/France/PAC-IHU-0633/2020|EPI_ISL_568904|20 ···················· ···················· ···················· ···················· ···················· ····················
K 5323 hCoV-19/France/PAC-IHU-0634/2020|EPI_ISL_568905|20 ···················· ···················· ···················· ···················· ···················· ····················
K 5324 hCoV-19/France/PAC-IHU-0635/2020|EPI_ISL_568906|20 ···················· ···················· ···················· ···················· ···················· ····················
K 5325 hCoV-19/France/PAC-IHU-0637/2020|EPI_ISL_568907|20 ···················· ··········A········· ···················· ···················· ···················· ····················
K 5326 hCoV-19/France/PAC-IHU-0638/2020|EPI_ISL_568908|20 ···················· ···················· ···················· ···················· ···················· ····················
K 5327 hCoV-19/France/PAC-IHU-0641/2020|EPI_ISL_568909|20 ···················· ···················· ···················· ···················· ···················· ····················
K 5328 hCoV-19/France/PAC-IHU-0646/2020|EPI_ISL_568910|20 ···················· ···················· ···················· ···················· ···················· ····················
K 5329 hCoV-19/France/PAC-IHU-0647/2020|EPI_ISL_568911|20 ···················· ···················· ···················· ···················· ···················· ····················
K 5330 hCoV-19/France/PAC-IHU-0648/2020|EPI_ISL_568912|20 ···················· ···················· ···················· ···················· ···················· ····················
K 5331 hCoV-19/France/PAC-IHU-0650/2020|EPI_ISL_568914|20 ···················· ···················· ···················· ···················· ···················· ····················
K 5332 hCoV-19/France/PAC-IHU-0651/2020|EPI_ISL_568915|20 ···················· ···················· ···················· ···················· ···················· ····················
K 5333 hCoV-19/France/PAC-IHU-0653/2020|EPI_ISL_568916|20 ···················· ···················· ···················· ···················· ···················· ····················
K 5334 hCoV-19/France/PAC-IHU-0655/2020|EPI_ISL_568917|20 ···················· ···················· ···················· ···················· ···················· ····················

| | | | | | | | | |
|---|---|---|---|---|---|---|---|---|
| K | 5335 | hCoV-19/France/PAC-IHU-0659/2020\|EPI_ISL_568918\|20 | ···················· | ···················· | ···················· | ···················· | ···················· | ···················· |
| K | 5336 | hCoV-19/France/PAC-IHU-0660/2020\|EPI_ISL_568919\|20 | ···················· | ···················· | ···················· | ···················· | ···················· | ···················· |
| K | 5337 | hCoV-19/France/PAC-IHU-0662/2020\|EPI_ISL_568920\|20 | ···················· | ···················· | ···················· | ···················· | ···················· | ···················· |
| K | 5338 | hCoV-19/France/PAC-IHU-0668/2020\|EPI_ISL_568921\|20 | ···················· | ···················· | ···················· | ···················· | ···················· | ···················· |
| K | 5339 | hCoV-19/France/PAC-IHU-0670/2020\|EPI_ISL_568922\|20 | ···················· | ···················· | ···················· | ···················· | ···················· | ···················· |
| K | 5340 | hCoV-19/France/PAC-IHU-0672/2020\|EPI_ISL_568924\|20 | ···················· | ···················· | ···················· | ···················· | ···················· | ···················· |
| K | 5341 | hCoV-19/France/PAC-IHU-0673/2020\|EPI_ISL_568925\|20 | ···················· | ···················· | ···················· | ···················· | ···················· | ···················· |
| K | 5342 | hCoV-19/France/PAC-IHU-0674/2020\|EPI_ISL_568926\|20 | ···················· | ···················· | ···················· | ···················· | ···················· | ···················· |
| K | 5343 | hCoV-19/France/PAC-IHU-0675/2020\|EPI_ISL_568927\|20 | ···················· | ···················· | ···················· | ···················· | ···················· | ···················· |
| K | 5344 | hCoV-19/France/PAC-IHU-0676/2020\|EPI_ISL_568928\|20 | ···················· | ···················· | ···················· | ···················· | ···················· | ···················· |
| K | 5345 | hCoV-19/France/PAC-IHU-0677/2020\|EPI_ISL_568929\|20 | ···················· | ···················· | ···················· | ···················· | ···················· | ···················· |
| K | 5346 | hCoV-19/France/PAC-IHU-0678/2020\|EPI_ISL_568930\|20 | ···················· | ···················· | ···················· | ···················· | ···················· | ···················· |
| K | 5347 | hCoV-19/France/PAC-IHU-0680/2020\|EPI_ISL_568931\|20 | ···················· | ···················· | ···················· | ···················· | ···················· | ···················· |
| K | 5348 | hCoV-19/France/PAC-IHU-0681/2020\|EPI_ISL_568932\|20 | ···················· | ···················· | ···················· | ···················· | ···················· | ···················· |
| K | 5349 | hCoV-19/France/PAC-IHU-0685/2020\|EPI_ISL_568933\|20 | ···················· | ···················· | ···················· | ···················· | ···················· | ···················· |
| K | 5351 | hCoV-19/France/PAC-IHU-0688/2020\|EPI_ISL_568935\|20 | ···················· | ···················· | ···················· | ···················· | ···················· | ···················· |
| K | 5352 | hCoV-19/France/PAC-IHU-0689/2020\|EPI_ISL_568936\|20 | ···················· | ···················· | ···················· | ···················· | ···················· | ···················· |
| K | 5353 | hCoV-19/France/PAC-IHU-0694/2020\|EPI_ISL_568939\|20 | ···················· | ···················· | ···················· | ···················· | ···················· | ···················· |
| K | 5354 | hCoV-19/France/PAC-IHU-0697/2020\|EPI_ISL_568940\|20 | ···················· | ···················· | ···················· | ···················· | ···················· | ···················· |
| K | 5355 | hCoV-19/France/PAC-IHU-0699/2020\|EPI_ISL_568941\|20 | ···················· | ···················· | ···················· | ···················· | ···················· | ···················· |
| K | 5356 | hCoV-19/France/PAC-IHU-0702/2020\|EPI_ISL_568942\|20 | ···················· | ···················· | ···················· | ···················· | ···················· | ···················· |
| K | 5357 | hCoV-19/France/PAC-IHU-0704/2020\|EPI_ISL_568944\|20 | ···················· | ···················· | ···················· | ···················· | ···················· | ·····N·············· |
| K | 5358 | hCoV-19/France/PAC-IHU-0708/2020\|EPI_ISL_568945\|20 | ···················· | ····T··············· | ···················· | ···················· | ···················· | ···················· |
| K | 5359 | hCoV-19/France/PAC-IHU-0726/2020\|EPI_ISL_568947\|20 | ···················· | ···················· | ···················· | ···················· | ···················· | ···················· |
| K | 5360 | hCoV-19/France/PAC-IHU-0728/2020\|EPI_ISL_568948\|20 | ···················· | ···················· | ···················· | ···················· | ···················· | ···················· |
| K | 5361 | hCoV-19/France/PAC-IHU-0729/2020\|EPI_ISL_568949\|20 | ···················· | ··········A········· | ···················· | ···················· | ···················· | ···················· |
| K | 5362 | hCoV-19/France/PAC-IHU-0730/2020\|EPI_ISL_568950\|20 | ···················· | ···················· | ···················· | ···················· | ···················· | ···················· |
| K | 5363 | hCoV-19/France/PAC-IHU-0731/2020\|EPI_ISL_568951\|20 | ···················· | ···················· | ···················· | ···················· | ···················· | ···················· |
| K | 5364 | hCoV-19/France/PAC-IHU-0733/2020\|EPI_ISL_568952\|20 | ···················· | ···················· | ···················· | ···················· | ···················· | ···················· |
| K | 5365 | hCoV-19/France/PAC-IHU-0736/2020\|EPI_ISL_568953\|20 | ···················· | ···················· | ···················· | ···················· | ···················· | ···················· |
| K | 5366 | hCoV-19/France/PAC-IHU-0739/2020\|EPI_ISL_568954\|20 | ···················· | ···················· | ···················· | ···················· | ···················· | ···················· |
| K | 5367 | hCoV-19/France/PAC-IHU-0743/2020\|EPI_ISL_568955\|20 | ···················· | ···················· | ···················· | ···················· | ···················· | ···················· |
| K | 5368 | hCoV-19/France/PAC-IHU-0756/2020\|EPI_ISL_568956\|20 | ···················· | ···················· | ···················· | ···················· | ···················· | ···················· |
| K | 5369 | hCoV-19/France/PAC-IHU-0781/2020\|EPI_ISL_568958\|20 | ···················· | ···················· | ···················· | ···················· | ···················· | ···················· |
| K | 5370 | hCoV-19/France/PAC-IHU-0783/2020\|EPI_ISL_568959\|20 | ···················· | ···················· | ···················· | ···················· | ···················· | ···················· |
| K | 5371 | hCoV-19/France/PAC-IHU-0784/2020\|EPI_ISL_568960\|20 | ···················· | ···················· | ···················· | ···················· | ···················· | ···················· |
| K | 5372 | hCoV-19/France/PAC-IHU-0785/2020\|EPI_ISL_568961\|20 | ···················· | ···················· | ···················· | ···················· | ···················· | ···················· |
| K | 5373 | hCoV-19/France/PAC-IHU-0786/2020\|EPI_ISL_568962\|20 | ···················· | ···················· | ···················· | ···················· | ···················· | ···················· |
| K | 5374 | hCoV-19/France/PAC-IHU-0787/2020\|EPI_ISL_568963\|20 | ···················· | ···················· | ···················· | ···················· | ···················· | ···················· |
| K | 5375 | hCoV-19/France/PAC-IHU-0788/2020\|EPI_ISL_568964\|20 | ···················· | ···················· | ···················· | ···················· | ···················· | ···················· |
| K | 5376 | hCoV-19/France/PAC-IHU-0789/2020\|EPI_ISL_568965\|20 | ···················· | ···················· | ···················· | ···················· | ···················· | ···················· |
| K | 5377 | hCoV-19/Gambia/NPHL-287/2020\|EPI_ISL_561300\|2020 | ···················· | ···················· | ···················· | ···················· | ···················· | ···················· |
| K | 5378 | hCoV-19/Gambia/NPHL-290/2020\|EPI_ISL_561303\|2020 | ···················· | ···················· | ···················· | ···················· | ···················· | ···················· |
| K | 5379 | hCoV-19/Gambia/NPHL-2909/2020\|EPI_ISL_561304\|2020- | ···················· | ···················· | ···················· | ···················· | ···················· | ···················· |
| K | 5380 | hCoV-19/Gambia/NPHL-295/2020\|EPI_ISL_561305\|2020 | ···················· | ···················· | ···················· | ···················· | ···················· | ···················· |
| K | 5381 | hCoV-19/Gambia/NPHL-297/2020\|EPI_ISL_561306\|2020 | ···················· | ···················· | ···················· | ···················· | ···················· | ···················· |
| K | 5382 | hCoV-19/Gambia/NPHL-2984/2020\|EPI_ISL_561307\|2020 | ···················· | ···················· | ···················· | ···················· | ···················· | ···················· |
| K | 5383 | hCoV-19/Gambia/NPHL-299/2020\|EPI_ISL_561308\|2020 | ···················· | ···················· | ···················· | ···················· | ···················· | ···················· |
| K | 5384 | hCoV-19/Gambia/NPHL-3047/2020\|EPI_ISL_561309\|2020- | ···················· | ···················· | ···················· | ···················· | ···················· | ···················· |
| K | 5385 | hCoV-19/Gambia/NPHL-748/2020\|EPI_ISL_561324\|2020 | ···················· | ···················· | ···················· | ···················· | ···················· | ···················· |
| K | 5386 | hCoV-19/Australia/VIC10480/2020\|EPI_ISL_562600\|202 | ···················· | ···················· | ···················· | ···················· | ···················· | ···················· |
| K | 5388 | hCoV-19/Australia/VIC10778/2020\|EPI_ISL_562602\|202 | ···················· | ···················· | ···················· | ···················· | ···················· | ···················· |
| K | 5389 | hCoV-19/Australia/VIC10922/2020\|EPI_ISL_562603\|202 | ···················· | ···················· | ···················· | ···················· | ···················· | ···················· |
| K | 5390 | hCoV-19/Australia/VIC10927/2020\|EPI_ISL_562604\|202 | ···················· | ···················· | ···················· | ···················· | ···················· | ···················· |
| K | 5391 | hCoV-19/Australia/VIC10930/2020\|EPI_ISL_562605\|202 | ···················· | ···················· | ···················· | ···················· | ···················· | ···················· |
| K | 5392 | hCoV-19/Australia/VIC11001/2020\|EPI_ISL_562606\|202 | ···················· | ···················· | ···················· | ···················· | ···················· | ···················· |
| K | 5393 | hCoV-19/Australia/VIC11027/2020\|EPI_ISL_562607\|202 | ···················· | ···················· | ···················· | ···················· | ···················· | ···················· |
| K | 5394 | hCoV-19/Australia/VIC11039/2020\|EPI_ISL_562608\|202 | ···················· | ···················· | ···················· | ···················· | ···················· | ···················· |
| K | 5395 | hCoV-19/Australia/VIC11062/2020\|EPI_ISL_562609\|202 | ···················· | ···················· | ···················· | ···················· | ···················· | ···················· |
| K | 5396 | hCoV-19/Australia/VIC11095/2020\|EPI_ISL_562610\|202 | ···················· | ···················· | ···················· | ···················· | ···················· | ···················· |
| K | 5397 | hCoV-19/Australia/VIC11167/2020\|EPI_ISL_562611\|202 | ···················· | ···················· | ···················· | ···················· | ···················· | ···················· |
| K | 5399 | hCoV-19/Australia/VIC11435/2020\|EPI_ISL_562613\|202 | ···················· | ···················· | ···················· | ···················· | ···················· | ···················· |
| K | 5402 | hCoV-19/Australia/VIC12793/2020\|EPI_ISL_562616\|202 | ···················· | ···················· | ···················· | ···················· | ···················· | ···················· |
| K | 5403 | hCoV-19/Australia/VIC13173/2020\|EPI_ISL_562617\|202 | ···················· | ···················· | ···················· | ···················· | ···················· | ···················· |
| K | 5404 | hCoV-19/Australia/VIC13621/2020\|EPI_ISL_562618\|202 | ···················· | ···················· | ···················· | ···················· | ···················· | ···················· |
| K | 5405 | hCoV-19/Australia/VIC13645/2020\|EPI_ISL_562619\|202 | ···················· | ···················· | ···················· | ···················· | ···················· | ···················· |
| K | 5407 | hCoV-19/Australia/VIC13772/2020\|EPI_ISL_562621\|202 | ···················· | ···················· | ···················· | ···················· | ···················· | ···················· |
| K | 5409 | hCoV-19/Australia/VIC7919/2020\|EPI_ISL_562623\|2020 | ···················· | ···················· | ···················· | ···················· | ···················· | ···················· |
| K | 5410 | hCoV-19/Australia/VIC8076/2020\|EPI_ISL_562624\|2020 | ···················· | ···················· | ···················· | ···················· | ···················· | ···················· |
| K | 5411 | hCoV-19/Australia/VIC8289/2020\|EPI_ISL_562625\|2020 | ···················· | ···················· | ···················· | ···················· | ···················· | ···················· |
| K | 5412 | hCoV-19/Australia/VIC8295/2020\|EPI_ISL_562626\|2020 | ···················· | ···················· | ···················· | ···················· | ···················· | ···················· |
| K | 5413 | hCoV-19/Australia/VIC8482/2020\|EPI_ISL_562627\|2020 | ···················· | ···················· | ···················· | ···················· | ···················· | ···················· |
| K | 5414 | hCoV-19/Australia/VIC8822/2020\|EPI_ISL_562628\|2020 | ···················· | ···················· | ···················· | ···················· | ···················· | ···················· |
| K | 5415 | hCoV-19/Australia/VIC9130/2020\|EPI_ISL_562629\|2020 | ···················· | ···················· | ···················· | ···················· | ···················· | ···················· |
| K | 5416 | hCoV-19/Australia/VIC9282/2020\|EPI_ISL_562630\|2020 | ···················· | ···················· | ···················· | ···················· | ···················· | ···················· |
| K | 5417 | hCoV-19/Australia/VIC9305/2020\|EPI_ISL_562631\|2020 | ···················· | ···················· | ···················· | ···················· | ···················· | ···················· |
| K | 5418 | hCoV-19/Australia/VIC9316/2020\|EPI_ISL_562632\|2020 | ···················· | ···················· | ···················· | ···················· | ···················· | ···················· |
| K | 5419 | hCoV-19/Australia/VIC9325/2020\|EPI_ISL_562633\|2020 | ···················· | ···················· | ···················· | ···················· | ···················· | ···················· |
| K | 5420 | hCoV-19/Australia/VIC9442/2020\|EPI_ISL_562634\|2020 | ···················· | ···················· | ···················· | ···················· | ···················· | ···················· |
| K | 5421 | hCoV-19/Australia/VIC9896/2020\|EPI_ISL_562635\|2020 | ···················· | ···················· | ···················· | ···················· | ···················· | ···················· |

| | | | | | | | | | |
|---|---|---|---|---|---|---|---|---|---|
| K | 5422 | hCoV-19/Australia/VIC9994/2020\|EPI_ISL_562636\|2020 | ···················· | ···················· | ···················· | ···················· | ···················· | ···················· | |
| K | 5423 | hCoV-19/Australia/VIC10023/2020\|EPI_ISL_562637\|202 | ···················· | ···················· | ···················· | ···················· | ···················· | ···················· | |
| K | 5425 | hCoV-19/Australia/VIC10634/2020\|EPI_ISL_562639\|202 | ···················· | ···················· | ···················· | ···················· | ···················· | ···················· | |
| K | 5426 | hCoV-19/Australia/VIC10894/2020\|EPI_ISL_562640\|202 | ···················· | ···················· | ···················· | ···················· | ···················· | ···················· | |
| K | 5429 | hCoV-19/Australia/VIC13928/2020\|EPI_ISL_562655\|202 | ···················· | ···················· | ···················· | ···················· | ···················· | ···················· | |
| K | 5430 | hCoV-19/Australia/VIC13987/2020\|EPI_ISL_563910\|202 | ···················· | ···················· | ···················· | ···················· | ···················· | ···················· | |
| K | 5432 | hCoV-19/Australia/VIC13989/2020\|EPI_ISL_563912\|202 | ···················· | ···················· | ···················· | ···················· | ···················· | ···················· | |
| K | 5434 | hCoV-19/Australia/VIC13992/2020\|EPI_ISL_563914\|202 | ···················· | ···················· | ···················· | ···················· | ···················· | ···················· | |
| K | 5436 | hCoV-19/Australia/VIC13995/2020\|EPI_ISL_563916\|202 | ···················· | ···················· | ···················· | ···················· | ···················· | ···················· | |
| K | 5437 | hCoV-19/Australia/VIC13997/2020\|EPI_ISL_563917\|202 | ···················· | ···················· | ···················· | ···················· | ···················· | ···················· | |
| K | 5439 | hCoV-19/Australia/VIC14001/2020\|EPI_ISL_563919\|202 | ···················· | ···················· | ···················· | ···················· | ···················· | ···················· | |
| K | 5440 | hCoV-19/Australia/VIC14002/2020\|EPI_ISL_563920\|202 | ···················· | ···················· | ···················· | ···················· | ···················· | ···················· | |
| K | 5441 | hCoV-19/Australia/VIC14009/2020\|EPI_ISL_563924\|202 | ···················· | ···················· | ···················· | ···················· | ···················· | ···················· | |
| K | 5442 | hCoV-19/Australia/VIC14015/2020\|EPI_ISL_563930\|202 | ···················· | ···················· | ···················· | ···················· | ···················· | ···················· | |
| K | 5446 | hCoV-19/Australia/VIC14023/2020\|EPI_ISL_563934\|202 | ···················· | ···················· | ···················· | ···················· | ···················· | ···················· | |
| K | 5450 | hCoV-19/Australia/VIC14031/2020\|EPI_ISL_563938\|202 | ···················· | ···················· | ···················· | ···················· | ···················· | ···················· | |
| K | 5454 | hCoV-19/Australia/VIC14041/2020\|EPI_ISL_563942\|202 | ···················· | ···················· | ···················· | ···················· | ···················· | ···················· | |
| K | 5457 | hCoV-19/Australia/VIC14047/2020\|EPI_ISL_563945\|202 | ···················· | ···················· | ···················· | ···················· | ···················· | ···················· | |
| K | 5459 | hCoV-19/Australia/VIC14056/2020\|EPI_ISL_563947\|202 | ···················· | ···················· | ···················· | ···················· | ···················· | ···················· | |
| K | 5460 | hCoV-19/Australia/VIC14057/2020\|EPI_ISL_563948\|202 | ···················· | ···················· | ···················· | ···················· | ···················· | ···················· | |
| K | 5463 | hCoV-19/Australia/VIC14061/2020\|EPI_ISL_563951\|202 | ···················· | ···················· | ···················· | ···················· | ···················· | ···················· | |
| K | 5464 | hCoV-19/Australia/VIC14063/2020\|EPI_ISL_563952\|202 | ···················· | ···················· | ···················· | ···················· | N··················· | ···················· | |
| K | 5465 | hCoV-19/Australia/VIC14068/2020\|EPI_ISL_563953\|202 | ···················· | ···················· | ···················· | ···················· | ···················· | ···················· | |
| K | 5466 | hCoV-19/Australia/VIC14071/2020\|EPI_ISL_563954\|202 | ···················· | ···················· | ···················· | ···················· | ···················· | ···················· | |
| K | 5468 | hCoV-19/Australia/VIC14074/2020\|EPI_ISL_563956\|202 | ···················· | ···················· | ···················· | ···················· | ···················· | ···················· | |
| K | 5469 | hCoV-19/Australia/VIC14075/2020\|EPI_ISL_563957\|202 | ···················· | ···················· | ···················· | ···················· | ···················· | ···················· | |
| K | 5472 | hCoV-19/Australia/VIC14081/2020\|EPI_ISL_563960\|202 | ···················· | ···················· | ···················· | ···················· | ···················· | ···················· | |
| K | 5477 | hCoV-19/Australia/VIC14100/2020\|EPI_ISL_563965\|202 | ···················· | ···················· | ···················· | ···················· | ···················· | ···················· | |
| K | 5478 | hCoV-19/Australia/VIC14101/2020\|EPI_ISL_563966\|202 | ···················· | ···················· | ···················· | ···················· | ···················· | ···················· | |
| K | 5479 | hCoV-19/Australia/VIC14102/2020\|EPI_ISL_563967\|202 | ···················· | ···················· | ···················· | ···················· | ···················· | ···················· | |
| K | 5480 | hCoV-19/Australia/VIC14103/2020\|EPI_ISL_563968\|202 | ···················· | ···················· | ···················· | ···················· | ···················· | ···················· | |
| K | 5481 | hCoV-19/Australia/VIC14104/2020\|EPI_ISL_563969\|202 | ···················· | ···················· | ···················· | ···················· | ···················· | ···················· | |
| K | 5482 | hCoV-19/Australia/VIC14106/2020\|EPI_ISL_563970\|202 | ···················· | ···················· | ···················· | ···················· | ···················· | ···················· | |
| K | 5483 | hCoV-19/Australia/VIC14109/2020\|EPI_ISL_563971\|202 | ···················· | ···················· | ···················· | ···················· | ···················· | ···················· | |
| K | 5484 | hCoV-19/Australia/VIC3034/2020\|EPI_ISL_563987\|2020 | ···················· | ···················· | ···················· | ···················· | ···················· | ···················· | |
| K | 5485 | hCoV-19/Australia/VIC3410/2020\|EPI_ISL_563988\|2020 | ···················· | ···················· | ···················· | ···················· | NNN················· | ···················· | 1 |
| K | 5486 | hCoV-19/Australia/VIC3983/2020\|EPI_ISL_563989\|2020 | ···················· | ···················· | ···················· | ···················· | ·NN················· | ···················· | |
| K | 5487 | hCoV-19/USA/GA-CDC-5118/2020\|EPI_ISL_566100\|2020-0 | ···················· | ····A··············· | ···················· | ···················· | ···················· | ···················· | |
| K | 5488 | hCoV-19/USA/GA-CDC-5119/2020\|EPI_ISL_566101\|2020-0 | ···················· | ····A··············· | ···················· | ···················· | ···················· | ···················· | |
| K | 5489 | hCoV-19/USA/GA-CDC-5122/2020\|EPI_ISL_566102\|2020-0 | ···················· | ····A··············· | ···················· | ···················· | ···················· | ···················· | |
| K | 5490 | hCoV-19/USA/GA-CDC-5123/2020\|EPI_ISL_566103\|2020-0 | ···················· | ····A··············· | ···················· | ···················· | ···················· | ···················· | |
| K | 5491 | hCoV-19/USA/GA-CDC-5124/2020\|EPI_ISL_566104\|2020-0 | ···················· | ····A··············· | ···················· | ···················· | ···················· | ···················· | |
| K | 5492 | hCoV-19/USA/GA-CDC-5125/2020\|EPI_ISL_566105\|2020-0 | ···················· | ····A··············· | ···················· | ···················· | ···················· | ···················· | |
| K | 5493 | hCoV-19/USA/GA-CDC-5126/2020\|EPI_ISL_566106\|2020-0 | ···················· | ····A··············· | ···················· | ···················· | ···················· | ···················· | |
| K | 5494 | hCoV-19/USA/GA-CDC-5127/2020\|EPI_ISL_566107\|2020-0 | ···················· | ····A··············· | ···················· | ···················· | ···················· | ···················· | |
| K | 5495 | hCoV-19/USA/DE-DHSS-F918682/2020\|EPI_ISL_566108\|20 | ···················· | ···················· | ···················· | ···················· | ···················· | ···················· | |
| K | 5496 | hCoV-19/England/ALDP-9CDC99/2020\|EPI_ISL_566111\|20 | ···················· | ···················· | ···················· | ···················· | ···················· | ···················· | |
| K | 5497 | hCoV-19/England/ALDP-9CDC6C/2020\|EPI_ISL_566113\|20 | ···················· | ···················· | ···················· | ···················· | ···················· | ···················· | |
| K | 5498 | hCoV-19/England/ALDP-9CCEC1/2020\|EPI_ISL_566114\|20 | ···················· | ·A·······A·········· | ···················· | ···················· | ···················· | ···················· | |
| K | 5499 | hCoV-19/England/ALDP-9CD90B/2020\|EPI_ISL_566115\|20 | ···················· | ···················· | ···················· | ···················· | ···················· | ···················· | |
| K | 5500 | hCoV-19/England/ALDP-9CE111/2020\|EPI_ISL_566116\|20 | ···················· | ···················· | ···················· | ···················· | ···················· | ···················· | |
| K | 5501 | hCoV-19/England/ALDP-9CD789/2020\|EPI_ISL_566117\|20 | ···················· | ···················· | ···················· | ···················· | ···················· | ···················· | |
| K | 5502 | hCoV-19/England/ALDP-9CD1B8/2020\|EPI_ISL_566118\|20 | ···················· | ···················· | ···················· | ···················· | ···················· | ···················· | |
| K | 5503 | hCoV-19/England/ALDP-9CCDA6/2020\|EPI_ISL_566119\|20 | ···················· | ···················· | ···················· | ···················· | ···················· | ···················· | |
| K | 5504 | hCoV-19/England/ALDP-9CD8FF/2020\|EPI_ISL_566120\|20 | ···················· | ···················· | ···················· | ···················· | ···················· | ···················· | |
| K | 5505 | hCoV-19/England/ALDP-9CDA9F/2020\|EPI_ISL_566121\|20 | ···················· | ···················· | ···················· | ···················· | ···················· | ···················· | |
| K | 5506 | hCoV-19/England/ALDP-9CD08E/2020\|EPI_ISL_566122\|20 | ···················· | ·A·······A·········· | ···················· | ···················· | ···················· | ···················· | |
| K | 5507 | hCoV-19/England/ALDP-9CD947/2020\|EPI_ISL_566123\|20 | ···················· | ···················· | ···················· | ···················· | ···················· | ···················· | |
| K | 5508 | hCoV-19/England/ALDP-9CDF45/2020\|EPI_ISL_566124\|20 | ···················· | ···················· | ···················· | ···················· | ···················· | ···················· | |
| K | 5509 | hCoV-19/England/ALDP-9CCE1C/2020\|EPI_ISL_566125\|20 | ···················· | ·A·················· | ···················· | ···················· | ···················· | ···A················ | |
| K | 5510 | hCoV-19/England/ALDP-9CDF09/2020\|EPI_ISL_566126\|20 | ···················· | ···················· | ···················· | ···················· | ···················· | ···················· | |
| K | 5511 | hCoV-19/England/ALDP-9CD66E/2020\|EPI_ISL_566127\|20 | ···················· | ·A·················· | ···················· | ···················· | ···················· | ···A················ | |
| K | 5512 | hCoV-19/England/ALDP-9CD32B/2020\|EPI_ISL_566128\|20 | ···················· | ···················· | ···················· | ···················· | ···················· | ···················· | |
| K | 5513 | hCoV-19/England/ALDP-9CCD79/2020\|EPI_ISL_566129\|20 | ···················· | ···················· | ···················· | ···················· | ···················· | ···················· | |
| K | 5514 | hCoV-19/England/ALDP-9CD358/2020\|EPI_ISL_566130\|20 | ···················· | ···················· | ···················· | ···················· | ···················· | ···················· | |
| K | 5515 | hCoV-19/England/ALDP-9CE06F/2020\|EPI_ISL_566131\|20 | ···················· | ·A·················· | ···················· | ···················· | ···················· | ···A················ | |
| K | 5516 | hCoV-19/England/ALDP-9CDB6F/2020\|EPI_ISL_566132\|20 | ···················· | ···················· | ···················· | ···················· | ···················· | ···················· | |
| K | 5517 | hCoV-19/England/ALDP-9CD130/2020\|EPI_ISL_566133\|20 | ···················· | ···················· | ···················· | ···················· | ···················· | ···················· | |
| K | 5518 | hCoV-19/England/ALDP-9CDC4E/2020\|EPI_ISL_566134\|20 | ···················· | ···················· | ···················· | ···················· | ······T············· | ···················· | |
| K | 5519 | hCoV-19/England/ALDP-9CD4EC/2020\|EPI_ISL_566135\|20 | ···················· | ·A·················· | ···················· | ···················· | ···················· | ···A················ | |
| K | 5520 | hCoV-19/England/ALDP-9CD81D/2020\|EPI_ISL_566136\|20 | ···················· | ···················· | ···················· | ···················· | ···················· | ···················· | |
| K | 5521 | hCoV-19/England/ALDP-9CCF82/2020\|EPI_ISL_566137\|20 | ···················· | ···················· | ···················· | ···················· | ···················· | ···················· | |
| K | 5522 | hCoV-19/England/ALDP-9CE005/2020\|EPI_ISL_566138\|20 | ···················· | ···················· | ···················· | ···················· | ···················· | ···················· | |
| K | 5523 | hCoV-19/England/ALDP-9CDDC3/2020\|EPI_ISL_566139\|20 | ···················· | ···················· | ···················· | ···················· | ···················· | ···················· | |
| K | 5524 | hCoV-19/England/ALDP-9CDDF0/2020\|EPI_ISL_566140\|20 | ···················· | ···················· | ···················· | ···················· | ···················· | ···················· | |
| K | 5525 | hCoV-19/England/ALDP-9CDCB7/2020\|EPI_ISL_566141\|20 | ···················· | ···················· | ···················· | ···················· | ···················· | ···················· | |
| K | 5526 | hCoV-19/England/ALDP-9CCD1F/2020\|EPI_ISL_566142\|20 | ···················· | ···················· | ···················· | ···················· | ···················· | ···················· | |
| K | 5527 | hCoV-19/England/ALDP-9CD992/2020\|EPI_ISL_566143\|20 | ···················· | ···················· | ···················· | ···················· | ···················· | ···················· | |
| K | 5528 | hCoV-19/England/ALDP-9CDFDC/2020\|EPI_ISL_566144\|20 | ···················· | ···················· | ···················· | ···················· | ···················· | ···················· | |
| K | 5529 | hCoV-19/England/ALDP-9CCF37/2020\|EPI_ISL_566145\|20 | ···················· | ···················· | ···················· | ···················· | ···················· | ···················· | |

| | | | | | | | | |
|---|---|---|---|---|---|---|---|---|
| K | 5530 | hCoV-19/England/ALDP-9CCD97/2020\|EPI_ISL_566146\|20 | ···················· | ···················· | ····················· | ···················· | ···················· | ···················· |
| K | 5531 | hCoV-19/England/ALDP-9CCFCE/2020\|EPI_ISL_566147\|20 | ···················· | ·A·················· | ····················· | ···················· | ···················· | ··A················· |
| K | 5532 | hCoV-19/England/ALDP-9CD552/2020\|EPI_ISL_566148\|20 | ···················· | ·A·················· | ····················· | ···················· | ···················· | ··A················· |
| K | 5533 | hCoV-19/England/ALDP-9CD07F/2020\|EPI_ISL_566149\|20 | ···················· | ·A·················· | ····················· | ···················· | ···················· | ··A················· |
| K | 5534 | hCoV-19/England/ALDP-9CD9DE/2020\|EPI_ISL_566190\|20 | ···················· | ···················· | ····················· | ···················· | ···················· | ···················· |
| K | 5535 | hCoV-19/England/ALDP-9CD18B/2020\|EPI_ISL_566191\|20 | ···················· | ···················· | ····················· | ···················· | ···················· | ···················· |
| K | 5536 | hCoV-19/England/ALDP-9CE050/2020\|EPI_ISL_566192\|20 | ···················· | ·A·················· | ····················· | ···················· | ···················· | ··A················· |
| K | 5537 | hCoV-19/England/ALDP-9CCD3D/2020\|EPI_ISL_566194\|20 | ···················· | ·A·················· | ····················· | ···················· | ···················· | ···················· |
| K | 5538 | hCoV-19/England/ALDP-9CE07E/2020\|EPI_ISL_566195\|20 | ···················· | ···················· | ····················· | ···················· | ···················· | ···················· |
| K | 5539 | hCoV-19/England/ALDP-9CD446/2020\|EPI_ISL_566196\|20 | ···················· | ···················· | ····················· | ···················· | ···················· | ···················· |
| K | 5540 | hCoV-19/England/ALDP-9CD437/2020\|EPI_ISL_566197\|20 | ···················· | ···················· | ····················· | ···················· | ···················· | ···················· |
| K | 5541 | hCoV-19/England/ALDP-9CD84A/2020\|EPI_ISL_566198\|20 | ···················· | ···················· | ····················· | ···················· | ···················· | ···················· |
| K | 5542 | hCoV-19/England/ALDP-9CD7B6/2020\|EPI_ISL_566199\|20 | ···················· | ···················· | ····················· | ···················· | ···················· | ···················· |
| K | 5543 | hCoV-19/England/QEUH-9C9D8B/2020\|EPI_ISL_567400\|20 | ···················· | ···················· | ····················· | ···················· | ···················· | ···················· |
| K | 5544 | hCoV-19/England/QEUH-9C9BDC/2020\|EPI_ISL_567401\|20 | ···················· | ···················· | ····················· | ···················· | ···················· | ···················· |
| K | 5545 | hCoV-19/England/QEUH-9C9DA9/2020\|EPI_ISL_567402\|20 | ···················· | ···················· | ····················· | ···················· | ···················· | ···················· |
| K | 5546 | hCoV-19/Wales/QEUH-9CA009/2020\|EPI_ISL_567403\|2020 | ···················· | ···················· | ····················· | ···················· | ···················· | ···················· |
| K | 5547 | hCoV-19/England/QEUH-9C9C24/2020\|EPI_ISL_567404\|20 | ···················· | ···················· | ····················· | ···················· | ···················· | ···················· |
| K | 5548 | hCoV-19/England/QEUH-9C92E6/2020\|EPI_ISL_567405\|20 | ···················· | ···················· | ····················· | ···················· | ···················· | ···················· |
| K | 5549 | hCoV-19/England/QEUH-9CA898/2020\|EPI_ISL_567406\|20 | ···················· | ···················· | ····················· | ···················· | ···················· | ···················· |
| K | 5550 | hCoV-19/England/QEUH-9C984E/2020\|EPI_ISL_567407\|20 | ···················· | ···················· | ····················· | ···················· | ···················· | ···················· |
| K | 5551 | hCoV-19/England/QEUH-9C99C3/2020\|EPI_ISL_567408\|20 | ···················· | ···················· | ····················· | ···················· | ···················· | ···················· |
| K | 5552 | hCoV-19/England/QEUH-9CA4EF/2020\|EPI_ISL_567409\|20 | ···················· | ···················· | ····················· | ···················· | ···················· | ···················· |
| K | 5553 | hCoV-19/England/QEUH-9C986C/2020\|EPI_ISL_567410\|20 | ···················· | ···················· | ····················· | ···················· | ···················· | ···················· |
| K | 5554 | hCoV-19/Wales/QEUH-9CA0BE/2020\|EPI_ISL_567411\|2020 | ···················· | ···················· | ····················· | ···················· | ···················· | ···················· |
| K | 5555 | hCoV-19/England/QEUH-9CA0FA/2020\|EPI_ISL_567412\|20 | ···················· | ···················· | ····················· | ···················· | ···················· | ···················· |
| K | 5556 | hCoV-19/England/QEUH-9CA0AF/2020\|EPI_ISL_567413\|20 | ···················· | ···················· | ····················· | ···················· | ···················· | ···················· |
| K | 5557 | hCoV-19/England/QEUH-9C93E3/2020\|EPI_ISL_567414\|20 | ···················· | ···················· | ····················· | ···················· | ···················· | ···················· |
| K | 5558 | hCoV-19/England/QEUH-9C9DC7/2020\|EPI_ISL_567415\|20 | ···················· | ···················· | ····················· | ···················· | ···················· | ···················· |
| K | 5559 | hCoV-19/England/QEUH-9CA054/2020\|EPI_ISL_567416\|20 | ···················· | ···················· | ····················· | ···················· | ···················· | ···················· |
| K | 5560 | hCoV-19/England/QEUH-9C9626/2020\|EPI_ISL_567417\|20 | ···················· | ·A·················· | ····················· | ···················· | ···················· | ··A················· |
| K | 5561 | hCoV-19/England/QEUH-9C933E/2020\|EPI_ISL_567418\|20 | ···················· | ···················· | ····················· | ···················· | ···················· | ···················· |
| K | 5562 | hCoV-19/England/QEUH-9CA3E2/2020\|EPI_ISL_567419\|20 | ···················· | ···················· | ····················· | ···················· | ···················· | ···················· |
| K | 5563 | hCoV-19/England/QEUH-9C96F9/2020\|EPI_ISL_567420\|20 | ···················· | ···················· | ····················· | ···················· | ···················· | ···················· |
| K | 5564 | hCoV-19/England/QEUH-9CA6E9/2020\|EPI_ISL_567421\|20 | ···················· | ···················· | ····················· | ···················· | ···················· | ···················· |
| K | 5565 | hCoV-19/England/QEUH-9CA212/2020\|EPI_ISL_567422\|20 | ···················· | ···················· | ····················· | ···················· | ···················· | ···················· |
| K | 5566 | hCoV-19/England/QEUH-9C9EE2/2020\|EPI_ISL_567423\|20 | ···················· | ···················· | ····················· | ···················· | ···················· | ···················· |
| K | 5567 | hCoV-19/England/QEUH-9C9E5B/2020\|EPI_ISL_567424\|20 | ···················· | ···················· | ····················· | ···················· | ···················· | ···················· |
| K | 5568 | hCoV-19/England/QEUH-9C9C60/2020\|EPI_ISL_567425\|20 | ···················· | ···················· | ····················· | ···················· | ······T············· | ···················· |
| K | 5569 | hCoV-19/England/QEUH-9CA75F/2020\|EPI_ISL_567426\|20 | ···················· | ···················· | ····················· | ···················· | ···················· | ···················· |
| K | 5570 | hCoV-19/England/QEUH-9C95B0/2020\|EPI_ISL_567427\|20 | ···················· | ···················· | ····················· | ···················· | ···················· | ···················· |
| K | 5571 | hCoV-19/England/QEUH-9CA78C/2020\|EPI_ISL_567428\|20 | ···················· | ···················· | ····················· | ···················· | ···················· | ···················· |
| K | 5572 | hCoV-19/England/QEUH-9C936B/2020\|EPI_ISL_567429\|20 | ···················· | ···················· | ····················· | ···················· | ···················· | ···················· |
| K | 5573 | hCoV-19/England/QEUH-9CA90E/2020\|EPI_ISL_567430\|20 | ···················· | ···················· | ····················· | ···················· | ···················· | ···················· |
| K | 5574 | hCoV-19/England/QEUH-9C9F2B/2020\|EPI_ISL_567431\|20 | ···················· | ···················· | ····················· | ···················· | ···················· | ···················· |
| K | 5575 | hCoV-19/England/QEUH-9C9547/2020\|EPI_ISL_567433\|20 | ···················· | ···················· | ····················· | ···················· | ···················· | ···················· |
| K | 5576 | hCoV-19/England/QEUH-9CA230/2020\|EPI_ISL_567434\|20 | ···················· | ···················· | ····················· | ···················· | ···················· | ···················· |
| K | 5577 | hCoV-19/England/QEUH-9C94E0/2020\|EPI_ISL_567435\|20 | ···················· | ···················· | ····················· | ···················· | ···················· | ···················· |
| K | 5578 | hCoV-19/England/QEUH-9C9BCD/2020\|EPI_ISL_567436\|20 | ···················· | ···················· | ····················· | ···················· | ···················· | ···················· |
| K | 5579 | hCoV-19/England/QEUH-9C9529/2020\|EPI_ISL_567437\|20 | ···················· | ···················· | ····················· | ···················· | ···················· | ···················· |
| K | 5580 | hCoV-19/England/QEUH-9C9DF4/2020\|EPI_ISL_567438\|20 | ···················· | ···················· | ····················· | ···················· | ···················· | ···················· |
| K | 5581 | hCoV-19/England/QEUH-9CA7E6/2020\|EPI_ISL_567439\|20 | ···················· | ···················· | ····················· | ···················· | ···················· | ···················· |
| K | 5582 | hCoV-19/England/QEUH-9C9A84/2020\|EPI_ISL_567440\|20 | ···················· | ···················· | ····················· | ···················· | ···················· | ···················· |
| K | 5583 | hCoV-19/England/QEUH-9CA7C8/2020\|EPI_ISL_567441\|20 | ···················· | ···················· | ····················· | ···················· | ···················· | ···················· |
| K | 5584 | hCoV-19/England/QEUH-9C944A/2020\|EPI_ISL_567442\|20 | ···················· | ···················· | ····················· | ···················· | ···················· | ···················· |
| K | 5585 | hCoV-19/England/QEUH-9C9486/2020\|EPI_ISL_567443\|20 | ···················· | ·A·················· | ····················· | ···················· | ···················· | ···················· |
| K | 5586 | hCoV-19/England/QEUH-9C94A4/2020\|EPI_ISL_567445\|20 | ···················· | ···················· | ····················· | ···················· | ···················· | ···················· |
| K | 5587 | hCoV-19/England/QEUH-9CA564/2020\|EPI_ISL_567446\|20 | ···················· | ···················· | ····················· | ···················· | ···················· | ···················· |
| K | 5588 | hCoV-19/England/QEUH-9C9AFD/2020\|EPI_ISL_567448\|20 | ···················· | ···················· | ····················· | ···················· | ···················· | ···················· |
| K | 5589 | hCoV-19/England/QEUH-9C9E4C/2020\|EPI_ISL_567449\|20 | ···················· | ···················· | ····················· | ···················· | ···················· | ···················· |
| K | 5590 | hCoV-19/England/QEUH-9CA5A0/2020\|EPI_ISL_567450\|20 | ···················· | ···················· | ····················· | ···················· | ···················· | ···················· |
| K | 5591 | hCoV-19/England/QEUH-9CA995/2020\|EPI_ISL_567451\|20 | ···················· | ·A·················· | ····················· | ···················· | ···················· | ···················· |
| K | 5592 | hCoV-19/England/QEUH-9CA27C/2020\|EPI_ISL_567452\|20 | ···················· | ···················· | ····················· | ···················· | ···················· | ···················· |
| K | 5593 | hCoV-19/England/QEUH-9C9583/2020\|EPI_ISL_567453\|20 | ···················· | ·A·················· | ····················· | ···················· | ···················· | ··A················· |
| K | 5594 | hCoV-19/England/QEUH-9CA160/2020\|EPI_ISL_567454\|20 | ···················· | ···················· | ····················· | ···················· | ···················· | ···················· |
| K | 5595 | hCoV-19/England/QEUH-9C97F6/2020\|EPI_ISL_567455\|20 | ···················· | ···················· | ····················· | ···················· | ···················· | ···················· |
| K | 5596 | hCoV-19/England/QEUH-9CA84D/2020\|EPI_ISL_567457\|20 | ···················· | ···················· | ····················· | ···················· | ···················· | ···················· |
| K | 5597 | hCoV-19/England/QEUH-9CA959/2020\|EPI_ISL_567458\|20 | ···················· | ···················· | ····················· | ···················· | ···················· | ···················· |
| K | 5598 | hCoV-19/England/QEUH-9C97D8/2020\|EPI_ISL_567459\|20 | ···················· | ···················· | ····················· | ···················· | ···················· | ···················· |
| K | 5599 | hCoV-19/England/QEUH-9CA6DA/2020\|EPI_ISL_567460\|20 | ···················· | ···················· | ····················· | ···················· | ···················· | ···················· |
| K | 5600 | hCoV-19/England/QEUH-9C9644/2020\|EPI_ISL_567461\|20 | ···················· | ···················· | ····················· | ···················· | ···················· | ···················· |
| K | 5601 | hCoV-19/England/QEUH-9C988A/2020\|EPI_ISL_567462\|20 | ···················· | ···················· | ····················· | ···················· | ···················· | ···················· |
| K | 5602 | hCoV-19/England/QEUH-9CA91D/2020\|EPI_ISL_567463\|20 | ···················· | ···················· | ····················· | ···················· | ···················· | ···················· |
| K | 5603 | hCoV-19/England/QEUH-9CA634/2020\|EPI_ISL_567465\|20 | ···················· | ···················· | ····················· | ···················· | ···················· | ···················· |
| K | 5604 | hCoV-19/England/QEUH-9C9F85/2020\|EPI_ISL_567466\|20 | ···················· | ·A·················· | ····················· | ···················· | ···················· | ··A················· |
| K | 5605 | hCoV-19/England/QEUH-9CA977/2020\|EPI_ISL_567467\|20 | ···················· | ···················· | ····················· | ···················· | ···················· | ···················· |
| K | 5606 | hCoV-19/England/QEUH-9C98E4/2020\|EPI_ISL_567468\|20 | ···················· | ·A·················· | ····················· | ···················· | ···················· | ···················· |
| K | 5607 | hCoV-19/England/QEUH-9CA045/2020\|EPI_ISL_567470\|20 | ···················· | ·A·················· | ····················· | ···················· | ···················· | ··A················· |
| K | 5608 | hCoV-19/England/QEUH-9CA731/2020\|EPI_ISL_567471\|20 | ···················· | ···················· | ····················· | ···················· | ···················· | ···················· |
| K | 5609 | hCoV-19/England/QEUH-9C9AEE/2020\|EPI_ISL_567472\|20 | ···················· | ···················· | ····················· | ···················· | ···················· | ···················· |

| | | | | | | | | |
|---|---|---|---|---|---|---|---|---|
| K | 5610 | hCoV-19/England/QEUH-9CA2A9/2020\|EPI_ISL_567473\|20 | ···················· | ···················· | ····················· | ···················· | ···················· | ···················· |
| K | 5611 | hCoV-19/England/QEUH-9CA8F2/2020\|EPI_ISL_567474\|20 | ···················· | ···················· | ····················· | ···················· | ···················· | ···················· |
| K | 5612 | hCoV-19/England/QEUH-9C9EF1/2020\|EPI_ISL_567475\|20 | ···················· | ·A·················· | ····················· | ···················· | ···················· | ··A················· |
| K | 5613 | hCoV-19/England/QEUH-9C99A5/2020\|EPI_ISL_567477\|20 | ···················· | ···················· | ····················· | ···················· | ···················· | ···················· |
| K | 5614 | hCoV-19/England/QEUH-9C94FF/2020\|EPI_ISL_567478\|20 | ···················· | ·A·······A·········· | ····················· | ···················· | ···················· | ···················· |
| K | 5615 | hCoV-19/England/QEUH-9C9E88/2020\|EPI_ISL_567479\|20 | ···················· | ···················· | ····················· | ···················· | ···················· | ···················· |
| K | 5616 | hCoV-19/England/QEUH-9CA063/2020\|EPI_ISL_567500\|20 | ···················· | ···················· | ····················· | ···················· | ···················· | ···················· |
| K | 5617 | hCoV-19/England/QEUH-9CA591/2020\|EPI_ISL_567501\|20 | ···················· | ···················· | ····················· | ···················· | ···················· | ···················· |
| K | 5618 | hCoV-19/England/QEUH-9C9398/2020\|EPI_ISL_567502\|20 | ···················· | ·A·················· | ····················· | ···················· | ···················· | ··A················· |
| K | 5619 | hCoV-19/England/QEUH-9C9B09/2020\|EPI_ISL_567503\|20 | ···················· | ···················· | ····················· | ···················· | ···················· | ···················· |
| K | 5620 | hCoV-19/England/QEUH-9C9301/2020\|EPI_ISL_567505\|20 | ···················· | ···················· | ····················· | ···················· | ···················· | ···················· |
| K | 5621 | hCoV-19/England/QEUH-9CA33D/2020\|EPI_ISL_567506\|20 | ···················· | ···················· | ····················· | ···················· | ···················· | ···················· |
| K | 5622 | hCoV-19/England/QEUH-9C98A8/2020\|EPI_ISL_567507\|20 | ···················· | ···················· | ····················· | ···················· | ···················· | ···················· |
| K | 5623 | hCoV-19/England/QEUH-9CA82F/2020\|EPI_ISL_567508\|20 | ···················· | ···················· | ····················· | ···················· | ···················· | ···················· |
| K | 5624 | hCoV-19/England/QEUH-9C9ADF/2020\|EPI_ISL_567509\|20 | ···················· | ···················· | ····················· | ···················· | ···················· | ···················· |
| K | 5625 | hCoV-19/England/QEUH-9C9ED3/2020\|EPI_ISL_567510\|20 | ···················· | ···················· | ····················· | ···················· | ···················· | ···················· |
| K | 5626 | hCoV-19/England/QEUH-9CA4D0/2020\|EPI_ISL_567511\|20 | ···················· | ···················· | ····················· | ···················· | ···················· | ···················· |
| K | 5627 | hCoV-19/England/QEUH-9CA2F4/2020\|EPI_ISL_567512\|20 | ···················· | ···················· | ····················· | ···················· | ···················· | ···················· |
| K | 5628 | hCoV-19/England/QEUH-9C9BBE/2020\|EPI_ISL_567513\|20 | ···················· | ·A·················· | ····················· | ···················· | ···················· | ··A················· |
| K | 5629 | hCoV-19/Kenya/C4214/2020\|EPI_ISL_568725\|2020-06-05 | ···················· | ···················· | ····················· | ···················· | ···················· | ···················· |
| K | 5630 | hCoV-19/Kenya/C4419/2020\|EPI_ISL_568727\|2020-05-07 | ···················· | ······A············· | ····················· | ···················· | ···················· | ···················· |
| K | 5631 | hCoV-19/Kenya/C4710/2020\|EPI_ISL_568735\|2020-05-08 | ···················· | ···················· | ····················· | ···················· | ···················· | ···················· |
| K | 5632 | hCoV-19/Kenya/C5384/2020\|EPI_ISL_568764\|2020-05-10 | ···················· | ···················· | ····················· | ···················· | ···················· | ···················· |
| K | 5633 | hCoV-19/Gambia/NPHL-3055/2020\|EPI_ISL_561310\|2020- | ···················· | ···················· | ····················· | ···················· | ···················· | ···················· |
| K | 5634 | hCoV-19/Gambia/NPHL-3062/2020\|EPI_ISL_561311\|2020 | ···················· | ···················· | ····················· | ···················· | ···················· | ···················· |
| K | 5635 | hCoV-19/Gambia/NPHL-3206/2020\|EPI_ISL_561313\|2020 | ···················· | ···················· | ····················· | ···················· | ···················· | ···················· |
| K | 5636 | hCoV-19/Gambia/NPHL-3247/2020\|EPI_ISL_561314\|2020 | ···················· | ···················· | ····················· | ···················· | ···················· | ···················· |
| K | 5637 | hCoV-19/Gambia/NPHL-3254/2020\|EPI_ISL_561315\|2020- | ···················· | ···················· | ····················· | ···················· | ···················· | ···················· |
| K | 5639 | hCoV-19/Gambia/NPHL-923/2020\|EPI_ISL_561330\|2020 | ···················· | ···················· | ····················· | ···················· | ···················· | ···················· |
| K | 5640 | hCoV-19/Gambia/NPHL-941/2020\|EPI_ISL_561332\|2020 | ···················· | ···················· | ····················· | ···················· | ···················· | ···················· |
| K | 5641 | hCoV-19/Gambia/NPHL-953/2020\|EPI_ISL_561333\|2020 | ···················· | ···················· | ····················· | ···················· | ···················· | ···················· |
| K | 5642 | hCoV-19/USA/DE-DHSS-F917656/2020\|EPI_ISL_561335\|20 | ···················· | ···················· | ····················· | ···················· | ···················· | ···················· |
| K | 5643 | hCoV-19/India/HR-IMT-BZ124/2020\|EPI_ISL_561339\|202 | ···················· | ·A·················· | ····················· | ···················· | ···················· | ··A················· |
| K | 5644 | hCoV-19/India/HR-IMT-BZ144/2020\|EPI_ISL_561340\|202 | ···················· | ·A·················· | ····················· | ···················· | ···················· | ··A················· |
| K | 5645 | hCoV-19/India/HR-IMT-BZ164/2020\|EPI_ISL_561341\|202 | ···················· | ·A·················· | ····················· | ···················· | ···················· | ··A················· |
| K | 5646 | hCoV-19/India/HR-IMT-CB126/2020\|EPI_ISL_561343\|202 | ···················· | ···················· | ····················· | ···················· | ···················· | ···················· |
| K | 5647 | hCoV-19/India/PB-IMT-L169/2020\|EPI_ISL_561344\|2020 | ···················· | ···················· | ····················· | ···················· | ···················· | ···················· |
| K | 5648 | hCoV-19/USA/DE-DHSS-F917718/2020\|EPI_ISL_561345\|20 | ···················· | ···················· | ····················· | ···················· | ···················· | ···················· |
| K | 5649 | hCoV-19/USA/DE-DHSS-F917736/2020\|EPI_ISL_561346\|20 | ···················· | ···················· | ····················· | ···················· | ···················· | ···················· |
| K | 5650 | hCoV-19/USA/DE-DHSS-F918676/2020\|EPI_ISL_561351\|20 | ···················· | ···················· | ····················· | ···················· | ···················· | ···················· |
| K | 5651 | hCoV-19/USA/DE-DHSS-F918677/2020\|EPI_ISL_561352\|20 | ···················· | ···················· | ····················· | ···················· | ···················· | ···················· |
| K | 5652 | hCoV-19/USA/DE-DHSS-F918750/2020\|EPI_ISL_561353\|20 | ···················· | ···················· | ····················· | ···················· | ···················· | ···················· |
| K | 5653 | hCoV-19/USA/DE-DHSS-F918898/2020\|EPI_ISL_561356\|20 | ···················· | ···················· | ····················· | ···················· | ···················· | ···················· |
| K | 5654 | hCoV-19/USA/DE-DHSS-F918926/2020\|EPI_ISL_561358\|20 | ···················· | ···················· | ····················· | ···················· | ···················· | ···················· |
| K | 5656 | hCoV-19/Spain/CN-IBV-97006522/2020\|EPI_ISL_561371\| | ···················· | ···················· | ····················· | ···················· | ···················· | N··················· |
| K | 5658 | hCoV-19/England/GSTT-4177/2020\|EPI_ISL_561373\|2020 | ···················· | ···················· | ····················· | ···················· | ···················· | ···················· |
| K | 5659 | hCoV-19/Australia/VIC11124/2020\|EPI_ISL_562641\|202 | ···················· | ···················· | ····················· | ···················· | ···················· | ···················· |
| K | 5661 | hCoV-19/Australia/VIC11436/2020\|EPI_ISL_562643\|202 | ···················· | ···················· | ····················· | ···················· | ···················· | ···················· |
| K | 5662 | hCoV-19/Australia/VIC11438/2020\|EPI_ISL_562644\|202 | ···················· | ···················· | ····················· | ···················· | ···················· | ···················· |
| K | 5663 | hCoV-19/Australia/VIC11562/2020\|EPI_ISL_562645\|202 | ···················· | ···················· | ····················· | ···················· | ···················· | ···················· |
| K | 5664 | hCoV-19/Australia/VIC11563/2020\|EPI_ISL_562646\|202 | ···················· | ···················· | ····················· | ···················· | ···················· | ···················· |
| K | 5665 | hCoV-19/Australia/VIC13368/2020\|EPI_ISL_562647\|202 | ···················· | ···················· | ····················· | ···················· | ···················· | ···················· |
| K | 5666 | hCoV-19/Australia/VIC13653/2020\|EPI_ISL_562648\|202 | ···················· | ···················· | ····················· | ···················· | ···················· | ···················· |
| K | 5667 | hCoV-19/Australia/VIC13666/2020\|EPI_ISL_562649\|202 | ···················· | ···················· | ····················· | ···················· | ···················· | ···················· |
| K | 5669 | hCoV-19/Australia/VIC13760/2020\|EPI_ISL_562651\|202 | ···················· | ···················· | ····················· | ···················· | ···················· | ···················· |
| K | 5671 | hCoV-19/Australia/VIC14007/2020\|EPI_ISL_562656\|202 | ···················· | ···················· | ····················· | ···················· | ···················· | ···················· |
| K | 5673 | hCoV-19/Australia/VIC7313/2020\|EPI_ISL_562658\|2020 | ···················· | ···················· | ····················· | ···················· | ···················· | ···················· |
| K | 5674 | hCoV-19/Australia/VIC7661/2020\|EPI_ISL_562659\|2020 | ···················· | ···················· | ····················· | ···················· | ···················· | ···················· |
| K | 5675 | hCoV-19/Australia/VIC7967/2020\|EPI_ISL_562660\|2020 | ···················· | ···················· | ····················· | ···················· | ···················· | ···················· |
| K | 5676 | hCoV-19/Australia/VIC8093/2020\|EPI_ISL_562661\|2020 | ···················· | ···················· | ····················· | ···················· | ···················· | ···················· |
| K | 5677 | hCoV-19/Australia/VIC8109/2020\|EPI_ISL_562662\|2020 | ···················· | ···················· | ····················· | ···················· | ···················· | ···················· |
| K | 5678 | hCoV-19/Australia/VIC8170/2020\|EPI_ISL_562663\|2020 | ···················· | ···················· | ····················· | ···················· | ···················· | ···················· |
| K | 5679 | hCoV-19/Australia/VIC8188/2020\|EPI_ISL_562664\|2020 | ···················· | ···················· | ····················· | ···················· | ···················· | ···················· |
| K | 5680 | hCoV-19/Australia/VIC8320/2020\|EPI_ISL_562665\|2020 | ···················· | ···················· | ····················· | ···················· | ···················· | ···················· |
| K | 5681 | hCoV-19/Australia/VIC8361/2020\|EPI_ISL_562666\|2020 | ···················· | ···················· | ····················· | ···················· | ···················· | ···················· |
| K | 5682 | hCoV-19/Australia/VIC8867/2020\|EPI_ISL_562667\|2020 | ···················· | ···················· | ····················· | ···················· | ···················· | ···················· |
| K | 5683 | hCoV-19/Australia/VIC8891/2020\|EPI_ISL_562668\|2020 | ···················· | ···················· | ····················· | ···················· | ···················· | ···················· |
| K | 5684 | hCoV-19/Australia/VIC9098/2020\|EPI_ISL_562669\|2020 | ···················· | ···················· | ···K················· | ···················· | ···················· | ···················· |
| K | 5685 | hCoV-19/Australia/VIC9259/2020\|EPI_ISL_562670\|2020 | ···················· | ···················· | ····················· | ···················· | ···················· | ···················· |
| K | 5686 | hCoV-19/Australia/VIC9262/2020\|EPI_ISL_562671\|2020 | ···················· | ···················· | ···K················· | ···················· | ···················· | ···················· |
| K | 5687 | hCoV-19/Australia/VIC9362/2020\|EPI_ISL_562672\|2020 | ···················· | ···················· | ····················· | ···················· | ···················· | ···················· |
| K | 5688 | hCoV-19/Australia/VIC9396/2020\|EPI_ISL_562673\|2020 | ···················· | ···················· | ····················· | ···················· | ···················· | ···················· |
| K | 5689 | hCoV-19/Australia/VIC9410/2020\|EPI_ISL_562674\|2020 | ···················· | ···················· | ····················· | ···················· | ···················· | ···················· |
| K | 5690 | hCoV-19/Australia/VIC9411/2020\|EPI_ISL_562675\|2020 | ···················· | ···················· | ····················· | ···················· | ···················· | ···················· |
| K | 5691 | hCoV-19/Australia/VIC9412/2020\|EPI_ISL_562676\|2020 | ···················· | ···················· | ····················· | ···················· | ···················· | ···················· |
| K | 5692 | hCoV-19/Australia/VIC9425/2020\|EPI_ISL_562677\|2020 | ···················· | ···················· | ····················· | ···················· | ···················· | ···················· |
| K | 5693 | hCoV-19/Australia/VIC9448/2020\|EPI_ISL_562678\|2020 | ···················· | ···················· | ····················· | ···················· | ···················· | ···················· |
| K | 5696 | hCoV-19/Australia/VIC9590/2020\|EPI_ISL_562681\|2020 | ···················· | ···················· | ····················· | ···················· | ···················· | ···················· |
| K | 5697 | hCoV-19/Australia/VIC9591/2020\|EPI_ISL_562682\|2020 | ···················· | ···················· | ····················· | ···················· | ···················· | ···················· |
| K | 5698 | hCoV-19/Australia/VIC9592/2020\|EPI_ISL_562683\|2020 | ···················· | ···················· | ····················· | ···················· | ···················· | ···················· |

| | | | | | | | |
|---|---|---|---|---|---|---|---|
| K | 5699 hCoV-19/Australia/VIC9593/2020\|EPI_ISL_562684\|2020 | ···················· | ···················· | ····················· | ···················· | ···················· | ···················· |
| K | 5700 hCoV-19/Australia/VIC9602/2020\|EPI_ISL_562685\|2020 | ···················· | ···················· | ····················· | ···················· | ···················· | ···················· |
| K | 5701 hCoV-19/Australia/VIC9868/2020\|EPI_ISL_562686\|2020 | ···················· | ···················· | ····················· | ···················· | ···················· | ···················· |
| K | 5702 hCoV-19/Australia/VIC9905/2020\|EPI_ISL_562687\|2020 | ···················· | ···················· | ····················· | ···················· | ···················· | ···················· |
| K | 5703 hCoV-19/Australia/VIC9247/2020\|EPI_ISL_562688\|2020 | ···················· | ···················· | ····················· | ···················· | ···················· | ···················· |
| K | 5704 hCoV-19/Australia/VIC10297/2020\|EPI_ISL_562691\|202 | ···················· | ···················· | ····················· | ···················· | ···················· | ···················· |
| K | 5705 hCoV-19/Australia/VIC10365/2020\|EPI_ISL_562692\|202 | ···················· | ···················· | ····················· | ···················· | ···················· | ···················· |
| K | 5708 hCoV-19/Australia/VIC11118/2020\|EPI_ISL_562695\|202 | ···················· | ···················· | ····················· | ···················· | ···················· | ···················· |
| K | 5711 hCoV-19/Australia/VIC9422/2020\|EPI_ISL_562698\|2020 | ···················· | ···················· | ····················· | ···················· | ···················· | ···················· |
| K | 5712 hCoV-19/Australia/VIC13658/2020\|EPI_ISL_563800\|202 | ···················· | ···················· | ····················· | ···················· | ···················· | ···················· |
| K | 5713 hCoV-19/Australia/VIC13660/2020\|EPI_ISL_563801\|202 | ···················· | ···················· | ····················· | ···················· | ···················· | ···················· |
| K | 5715 hCoV-19/Australia/VIC13669/2020\|EPI_ISL_563803\|202 | ···················· | ····T··············· | ····················· | ···················· | ···················· | ···················· |
| K | 5716 hCoV-19/Australia/VIC13670/2020\|EPI_ISL_563804\|202 | ···················· | ···················· | ····················· | ···················· | ···················· | ···················· |
| K | 5717 hCoV-19/Australia/VIC13671/2020\|EPI_ISL_563805\|202 | ···················· | ···················· | ····················· | ···················· | ···················· | ···················· |
| K | 5719 hCoV-19/Australia/VIC13674/2020\|EPI_ISL_563807\|202 | ···················· | ···················· | ····················· | ···················· | ···················· | ···················· |
| K | 5721 hCoV-19/Australia/VIC13699/2020\|EPI_ISL_563809\|202 | ···················· | ···················· | ····················· | ···················· | ···················· | ···················· |
| K | 5722 hCoV-19/Australia/VIC13700/2020\|EPI_ISL_563810\|202 | ···················· | ···················· | ····················· | ···················· | ···················· | ···················· |
| K | 5723 hCoV-19/Australia/VIC13705/2020\|EPI_ISL_563811\|202 | ···················· | ···················· | ····················· | ···················· | ···················· | ···················· |
| K | 5724 hCoV-19/Australia/VIC13708/2020\|EPI_ISL_563812\|202 | ···················· | ···················· | ····················· | ···················· | ···················· | ···················· |
| K | 5725 hCoV-19/Australia/VIC13714/2020\|EPI_ISL_563813\|202 | ···················· | ···················· | ····················· | ···················· | ···················· | ···················· |
| K | 5727 hCoV-19/Australia/VIC13717/2020\|EPI_ISL_563815\|202 | ···················· | ···················· | ····················· | ···················· | ···················· | ···················· |
| K | 5728 hCoV-19/Australia/VIC13720/2020\|EPI_ISL_563816\|202 | ···················· | ···················· | ····················· | ···················· | ···················· | ···················· |
| K | 5729 hCoV-19/Australia/VIC13728/2020\|EPI_ISL_563817\|202 | ···················· | ···················· | ····················· | ···················· | ···················· | ···················· |
| K | 5730 hCoV-19/Australia/VIC13734/2020\|EPI_ISL_563818\|202 | ···················· | ···················· | ····················· | ···················· | ···················· | ···················· |
| K | 5732 hCoV-19/Australia/VIC13741/2020\|EPI_ISL_563820\|202 | ···················· | ···················· | ····················· | ···················· | ···················· | ···················· |
| K | 5733 hCoV-19/Australia/VIC13743/2020\|EPI_ISL_563821\|202 | ···················· | ···················· | ····················· | ···················· | ···················· | ···················· |
| K | 5735 hCoV-19/Australia/VIC13748/2020\|EPI_ISL_563823\|202 | ···················· | ···················· | ····················· | ···················· | ···················· | ···················· |
| K | 5737 hCoV-19/Australia/VIC13753/2020\|EPI_ISL_563825\|202 | ···················· | ···················· | ····················· | ···················· | ···················· | ···················· |
| K | 5738 hCoV-19/Australia/VIC13755/2020\|EPI_ISL_563826\|202 | ···················· | ···················· | ····················· | ···················· | ···················· | ···················· |
| K | 5739 hCoV-19/Australia/VIC13757/2020\|EPI_ISL_563827\|202 | ···················· | ···················· | ····················· | ···················· | ···················· | ···················· |
| K | 5740 hCoV-19/Australia/VIC13762/2020\|EPI_ISL_563828\|202 | ···················· | ···················· | ····················· | ···················· | ···················· | ···················· |
| K | 5741 hCoV-19/Australia/VIC13764/2020\|EPI_ISL_563829\|202 | ···················· | ···················· | ····················· | ···················· | ···················· | ···················· |
| K | 5743 hCoV-19/Australia/VIC14114/2020\|EPI_ISL_563973\|202 | ···················· | ···················· | ····················· | ···················· | ···················· | ···················· |
| K | 5744 hCoV-19/Australia/VIC14115/2020\|EPI_ISL_563974\|202 | ···················· | ···················· | ····················· | ···················· | ···················· | ···················· |
| K | 5745 hCoV-19/Australia/VIC14116/2020\|EPI_ISL_563975\|202 | ···················· | ···················· | ····················· | ···················· | ···················· | ···················· |
| K | 5749 hCoV-19/Australia/VIC2072/2020\|EPI_ISL_563979\|2020 | ···················· | ···················· | ····················· | ···················· | ···················· | ···················· |
| K | 5757 hCoV-19/Australia/VIC3984/2020\|EPI_ISL_563990\|2020 | ···················· | ···················· | ····················· | ···················· | ···················· | ···················· |
| K | 5758 hCoV-19/Australia/VIC6379/2020\|EPI_ISL_563991\|2020 | ···················· | ···················· | ····················· | ···················· | ···················· | ···················· |
| K | 5759 hCoV-19/Australia/VIC6491/2020\|EPI_ISL_563992\|2020 | ···················· | ···················· | ····················· | ···················· | ···················· | ···················· |
| K | 5760 hCoV-19/Australia/VIC6493/2020\|EPI_ISL_563993\|2020 | ···················· | ···················· | ····················· | ···················· | ···················· | ···················· |
| K | 5761 hCoV-19/Australia/VIC6494/2020\|EPI_ISL_563994\|2020 | ···················· | ···················· | ····················· | ···················· | ···················· | ···················· |
| K | 5762 hCoV-19/Australia/VIC6513/2020\|EPI_ISL_563995\|2020 | ···················· | ···················· | ····················· | ···················· | ···················· | ···················· |
| K | 5763 hCoV-19/Australia/VIC6514/2020\|EPI_ISL_563996\|2020 | ···················· | ···················· | ····················· | ···················· | ···················· | ···················· |
| K | 5764 hCoV-19/Australia/VIC6694/2020\|EPI_ISL_563997\|2020 | ···················· | ···················· | ···K················· | ···················· | ···················· | ···················· |
| K | 5765 hCoV-19/Australia/VIC6696/2020\|EPI_ISL_563998\|2020 | ···················· | ···················· | ····················· | ···················· | ···················· | ···················· |
| K | 5766 hCoV-19/Australia/VIC6697/2020\|EPI_ISL_563999\|2020 | ···················· | ···················· | ····················· | ···················· | ···················· | ···················· |
| K | 5767 hCoV-19/USA/MI-MDHHS-SC21979/2020\|EPI_ISL_566029\|2 | ···················· | ···················· | ····················· | ···················· | ···················· | ···················· |
| K | 5768 hCoV-19/USA/MI-MDHHS-SC21980/2020\|EPI_ISL_566030\|2 | ···················· | ···················· | ····················· | ···················· | ···················· | ···················· |
| K | 5769 hCoV-19/USA/MI-MDHHS-SC21981/2020\|EPI_ISL_566031\|2 | ···················· | ··············C····· | ····················· | ···················· | ···················· | ···················· |
| K | 5770 hCoV-19/USA/MI-MDHHS-SC21982/2020\|EPI_ISL_566032\|2 | ···················· | ·············A······ | ····················· | ···················· | ···················· | ···················· |
| K | 5771 hCoV-19/USA/MI-MDHHS-SC21983/2020\|EPI_ISL_566033\|2 | ···················· | ···················· | ····················· | ···················· | ···················· | ···················· |
| K | 5772 hCoV-19/USA/MI-MDHHS-SC21984/2020\|EPI_ISL_566034\|2 | ···················· | ···················· | ····················· | ···················· | ···················· | ···················· |
| K | 5773 hCoV-19/USA/MI-MDHHS-SC21985/2020\|EPI_ISL_566035\|2 | ···················· | ···················· | ····················· | ···················· | ···················· | ···················· |
| K | 5774 hCoV-19/England/201090128/2020\|EPI_ISL_566047\|2020 | ···················· | ···················· | ····················· | ···················· | ···················· | ···················· |
| K | 5775 hCoV-19/England/201090129/2020\|EPI_ISL_566048\|2020 | ···················· | ···················· | ····················· | ···················· | ···················· | ···················· |
| K | 5776 hCoV-19/England/201100832/2020\|EPI_ISL_566049\|2020 | ···················· | ···················· | ····················· | ···················· | ···················· | ···················· |
| K | 5777 hCoV-19/England/202320700/2020\|EPI_ISL_566050\|2020 | ···················· | ···················· | ····················· | ···················· | ···················· | ···················· |
| K | 5778 hCoV-19/England/202322095/2020\|EPI_ISL_566051\|2020 | ···················· | ···················· | ····················· | ···················· | ···················· | ···················· |
| K | 5779 hCoV-19/England/202540896/2020\|EPI_ISL_566052\|2020 | ···················· | ···················· | ····················· | ···················· | ···················· | ···················· |
| K | 5780 hCoV-19/England/202580750/2020\|EPI_ISL_566053\|2020 | ···················· | ···················· | ····················· | ···················· | ···················· | ···················· |
| K | 5781 hCoV-19/England/202600045/2020\|EPI_ISL_566054\|2020 | ···················· | ···················· | ····················· | ···················· | ···················· | ···················· |
| K | 5782 hCoV-19/England/202620695/2020\|EPI_ISL_566055\|2020 | ···················· | ···················· | ····················· | ···················· | ···················· | ···················· |
| K | 5783 hCoV-19/England/202640035/2020\|EPI_ISL_566056\|2020 | ···················· | ···················· | ····················· | ···················· | ···················· | ···················· |
| K | 5784 hCoV-19/England/202660056/2020\|EPI_ISL_566057\|2020 | ···················· | ···················· | ····················· | ···················· | ···················· | ···················· |
| K | 5785 hCoV-19/England/202740182/2020\|EPI_ISL_566058\|2020 | ···················· | ···················· | ····················· | ···················· | ···················· | ···················· |
| K | 5786 hCoV-19/England/202760355/2020\|EPI_ISL_566059\|2020 | ···················· | ···················· | ····················· | ···················· | ···················· | ···················· |
| K | 5787 hCoV-19/England/202840488/2020\|EPI_ISL_566060\|2020 | ···················· | ···················· | ····················· | ···················· | ···················· | ···················· |
| K | 5788 hCoV-19/England/202880975/2020\|EPI_ISL_566061\|2020 | ···················· | ···················· | ····················· | ···················· | ···················· | ···················· |
| K | 5789 hCoV-19/England/202890228/2020\|EPI_ISL_566062\|2020 | ···················· | ·A·················· | ····················· | ···················· | ···················· | ···················· |
| K | 5790 hCoV-19/England/203060269/2020\|EPI_ISL_566063\|2020 | ···················· | ···················· | ····················· | ···················· | ···················· | ···················· |
| K | 5791 hCoV-19/England/203180881/2020\|EPI_ISL_566064\|2020 | ···················· | ···················· | ····················· | ···················· | ···················· | ···················· |
| K | 5792 hCoV-19/England/203260119/2020\|EPI_ISL_566065\|2020 | ···················· | ···················· | ····················· | ···················· | ···················· | ···················· |
| K | 5793 hCoV-19/England/203420511/2020\|EPI_ISL_566066\|2020 | ···················· | ·A·················· | ····················· | ···················· | ···················· | ···················· |
| K | 5794 hCoV-19/England/203440490/2020\|EPI_ISL_566067\|2020 | ···················· | ···················· | ····················· | ···················· | ···················· | ···················· |
| K | 5795 hCoV-19/England/203480317/2020\|EPI_ISL_566068\|2020 | ···················· | ···················· | ····················· | ···················· | ···················· | ···················· |
| K | 5796 hCoV-19/England/203580245/2020\|EPI_ISL_566069\|2020 | ···················· | ···················· | ····················· | ···················· | ···················· | ···················· |
| K | 5797 hCoV-19/England/203580247/2020\|EPI_ISL_566070\|2020 | ···················· | ···················· | ····················· | ···················· | ···················· | ···················· |
| K | 5798 hCoV-19/England/203580248/2020\|EPI_ISL_566071\|2020 | ···················· | ···················· | ····················· | ···················· | ···················· | ···················· |
| K | 5799 hCoV-19/England/203580286/2020\|EPI_ISL_566072\|2020 | ···················· | ···················· | ····················· | ···················· | ···················· | ···················· |
| K | 5800 hCoV-19/England/203590010/2020\|EPI_ISL_566073\|2020 | ···················· | ···················· | ····················· | ···················· | ···················· | ···················· |

K 5801 hCoV-19/England/203660640/2020|EPI_ISL_566074|2020 ···················· ···················· ···················· ···················· ···················· ····················
K 5802 hCoV-19/Russia/MOW-RII-MH249S/2020|EPI_ISL_566077| ···················· ···················· ···················· ···················· ···················· ····················
K 5803 hCoV-19/Russia/MOW-RII-MH250S/2020|EPI_ISL_566078| ···················· ···················· ···················· ···················· ···················· ····················
K 5804 hCoV-19/Russia/MOW-RII-MH251S/2020|EPI_ISL_566079| ···················· ···················· ···················· ···················· ···················· ····················
K 5805 hCoV-19/Russia/MOW-RII-MH252S/2020|EPI_ISL_566080| ···················· ···················· ···················· ···················· ···················· ····················
K 5806 hCoV-19/Russia/MOW-RII-MH254S/2020|EPI_ISL_566081| ···················· ···················· ···················· ···················· ···················· ····················
K 5807 hCoV-19/Russia/MOW-RII-MH255S/2020|EPI_ISL_566082| ···················· ···················· ···················· ···················· ···················· ····················
K 5808 hCoV-19/Russia/MOW-RII-MH256S/2020|EPI_ISL_566083| ···················· ···················· ···················· ···················· ···················· ····················
K 5809 hCoV-19/USA/DE-DHSS-F918573/2020|EPI_ISL_566084|20 ···················· ···················· ···················· ···················· ···················· ····················
K 5810 hCoV-19/USA/GA-CDC-5095/2020|EPI_ISL_566085|2020-0 ···················· ····A··············· ···················· ···················· ···················· ····················
K 5811 hCoV-19/USA/GA-CDC-5097/2020|EPI_ISL_566086|2020-0 ···················· ····A··············· ···················· ···················· ···················· ····················
K 5812 hCoV-19/USA/GA-CDC-5099/2020|EPI_ISL_566087|2020-0 ···················· ····A··············· ···················· ···················· ···················· ····················
K 5813 hCoV-19/USA/GA-CDC-5100/2020|EPI_ISL_566088|2020-0 ···················· ····A··············· ···················· ···················· ···················· ····················
K 5814 hCoV-19/USA/GA-CDC-5101/2020|EPI_ISL_566089|2020-0 ···················· ····A··············· ···················· ···················· ···················· ····················
K 5815 hCoV-19/USA/GA-CDC-5102/2020|EPI_ISL_566090|2020-0 ···················· ····A··············· ···················· ···················· ···················· ····················
K 5816 hCoV-19/USA/GA-CDC-5103/2020|EPI_ISL_566091|2020-0 ···················· ····A··············· ···················· ···················· ···················· ····················
K 5817 hCoV-19/USA/GA-CDC-5104/2020|EPI_ISL_566092|2020-0 ···················· ····A··············· ···················· ···················· ···················· ····················
K 5818 hCoV-19/USA/GA-CDC-5107/2020|EPI_ISL_566093|2020-0 ···················· ····A··············· ···················· ···················· ···················· ····················
K 5819 hCoV-19/USA/GA-CDC-5108/2020|EPI_ISL_566094|2020-0 ···················· ····A··············· ···················· ···················· ···················· ····················
K 5820 hCoV-19/USA/GA-CDC-5110/2020|EPI_ISL_566095|2020-0 ···················· ····A··············· ···················· ···················· ···················· ····················
K 5821 hCoV-19/USA/GA-CDC-5112/2020|EPI_ISL_566096|2020-0 ···················· ····A··············· ···················· ···················· ···················· ····················
K 5822 hCoV-19/USA/GA-CDC-5113/2020|EPI_ISL_566097|2020-0 ···················· ····A··············· ···················· ···················· ···················· ····················
K 5823 hCoV-19/USA/GA-CDC-5115/2020|EPI_ISL_566098|2020-0 ···················· ····A··············· ···················· ···················· ···················· ····················
K 5824 hCoV-19/USA/GA-CDC-5117/2020|EPI_ISL_566099|2020-0 ···················· ····A··············· ···················· ···················· ···················· ····················
K 5825 hCoV-19/England/QEUH-9CA41C/2020|EPI_ISL_567360|20 ···················· ···················· ···················· ···················· ···················· ····················
K 5826 hCoV-19/England/QEUH-9CA081/2020|EPI_ISL_567361|20 ···················· ···················· ···················· ···················· ···················· ····················
K 5827 hCoV-19/England/QEUH-9CA36A/2020|EPI_ISL_567362|20 ···················· ···················· ···················· ···················· ···················· ····················
K 5828 hCoV-19/England/QEUH-9C96DB/2020|EPI_ISL_567363|20 ···················· ···················· ···················· ···················· ···················· ····················
K 5829 hCoV-19/England/QEUH-9C935C/2020|EPI_ISL_567364|20 ···················· ···················· ···················· ···················· ···················· ····················
K 5830 hCoV-19/England/QEUH-9CA8B6/2020|EPI_ISL_567365|20 ···················· ···················· ···················· ···················· ···················· ····················
K 5831 hCoV-19/England/QEUH-9CA85C/2020|EPI_ISL_567366|20 ···················· ···················· ···················· ···················· ···················· ····················
K 5832 hCoV-19/England/QEUH-9C9FD0/2020|EPI_ISL_567367|20 ···················· ···················· ···················· ···················· ···················· ····················
K 5833 hCoV-19/Wales/QEUH-9CA0CD/2020|EPI_ISL_567368|2020 ···················· ···················· ···················· ···················· ···················· ····················
K 5834 hCoV-19/England/QEUH-9C9BEB/2020|EPI_ISL_567369|20 ···················· ···················· ···················· ···················· ···················· ····················
K 5835 hCoV-19/England/QEUH-9CA8A7/2020|EPI_ISL_567370|20 ···················· ···················· ···················· ···················· ···················· ····················
K 5836 hCoV-19/England/QEUH-9C9E3D/2020|EPI_ISL_567372|20 ···················· ·A·················· ···················· ···················· ···················· ··A·················
K 5837 hCoV-19/England/QEUH-9C9732/2020|EPI_ISL_567373|20 ···················· ···················· ···················· ···················· ···················· ····················
K 5838 hCoV-19/England/QEUH-9C9D5E/2020|EPI_ISL_567374|20 ···················· ···················· ···················· ···················· ···················· ····················
K 5839 hCoV-19/England/QEUH-9C9714/2020|EPI_ISL_567376|20 ···················· ···················· ···················· ···················· ···················· ····················
K 5840 hCoV-19/England/QEUH-9C95ED/2020|EPI_ISL_567377|20 ···················· ···················· ···················· ···················· ···················· ····················
K 5841 hCoV-19/England/QEUH-9C9C15/2020|EPI_ISL_567378|20 ···················· ···················· ···················· ···················· ···················· ····················
K 5842 hCoV-19/England/QEUH-9C9CBB/2020|EPI_ISL_567379|20 ···················· ···················· ···················· ···················· ···················· ····················
K 5843 hCoV-19/England/QEUH-9C993C/2020|EPI_ISL_567380|20 ···················· ···················· ···················· ···················· ···················· ····················
K 5844 hCoV-19/England/QEUH-9C9565/2020|EPI_ISL_567381|20 ···················· ···················· ···················· ···················· ···················· ····················
K 5845 hCoV-19/England/QEUH-9CA5DD/2020|EPI_ISL_567382|20 ···················· ·A·················· ···················· ···················· ···················· ····················
K 5846 hCoV-19/England/QEUH-9CA458/2020|EPI_ISL_567383|20 ···················· ···················· ···················· ···················· ···················· ····················
K 5847 hCoV-19/England/QEUH-9CA740/2020|EPI_ISL_567384|20 ···················· ···················· ···················· ···················· ······T············· ····················
K 5848 hCoV-19/England/QEUH-9C9E6A/2020|EPI_ISL_567386|20 ···················· ···················· ···················· ···················· ···················· ····················
K 5849 hCoV-19/England/QEUH-9CA2E5/2020|EPI_ISL_567388|20 ···················· ···················· ···················· ···················· ···················· ····················
K 5850 hCoV-19/England/QEUH-9CA4FE/2020|EPI_ISL_567389|20 ···················· ···················· ···················· ···················· ····T··············· ····················
K 5851 hCoV-19/England/QEUH-9CA5EC/2020|EPI_ISL_567391|20 ···················· ······C············· ···················· ···················· ···················· ····················
K 5852 hCoV-19/England/QEUH-9CA300/2020|EPI_ISL_567392|20 ···················· ···················· ···················· ···················· ···················· ····················
K 5853 hCoV-19/England/QEUH-9CA7B9/2020|EPI_ISL_567393|20 ···················· ···················· ···················· ···················· ···················· ····················
K 5854 hCoV-19/England/QEUH-9CA94A/2020|EPI_ISL_567394|20 ···················· ···················· ···················· ···················· ···················· ····················
K 5855 hCoV-19/Wales/QEUH-9C951A/2020|EPI_ISL_567396|2020 ···················· ···················· ···················· ···················· ···················· ····················
K 5856 hCoV-19/England/QEUH-9C9E00/2020|EPI_ISL_567397|20 ···················· ···················· ···················· ···················· ···················· ····················
K 5857 hCoV-19/Wales/QEUH-9C9EC4/2020|EPI_ISL_567398|2020 ···················· ···················· ···················· ···················· ···················· ····················
K 5858 hCoV-19/England/QEUH-9CA036/2020|EPI_ISL_567399|20 ···················· ···················· ···················· ···················· ···················· ····················
K 5860 hCoV-19/USA/FL-BPHL-1519/2020|EPI_ISL_568617|2020- ···················· ···················· ···················· ···················· ···················· ····················
K 5861 hCoV-19/USA/FL-BPHL-1520/2020|EPI_ISL_568618|2020- ···················· ···················· ···················· ···················· ···················· ····················
K 5862 hCoV-19/USA/FL-BPHL-1521/2020|EPI_ISL_568619|2020- ···················· ····A··············· ···················· ···················· ···················· ····················
K 5863 hCoV-19/USA/FL-BPHL-1522/2020|EPI_ISL_568620|2020- ···················· ···················· ···················· ···················· ···················· ····················
K 5864 hCoV-19/USA/FL-BPHL-1523/2020|EPI_ISL_568621|2020- ···················· ···················· ···················· ···················· ···················· ····················
K 5866 hCoV-19/USA/FL-BPHL-1525/2020|EPI_ISL_568623|2020- ···················· ···················· ···················· ···················· ···················· ····················
K 5867 hCoV-19/USA/FL-BPHL-1526/2020|EPI_ISL_568624|2020- ···················· ···················· ···················· ···················· ···················· ····················
K 5868 hCoV-19/Indonesia/JK-EIJK-39/2020|EPI_ISL_568690|2 ···················· ···················· ···················· ···················· ···················· ····················
K 5869 hCoV-19/Indonesia/JK-EIJK-40/2020|EPI_ISL_568691|2 ···················· ···················· ···················· ···················· ···················· ····················
K 5870 hCoV-19/Indonesia/JK-EIJK-43/2020|EPI_ISL_568692|2 ···················· ···················· ···················· ···················· ···················· ····················
K 5871 hCoV-19/Indonesia/PP-EIJK-11/2020|EPI_ISL_568693|2 ···················· ···················· ···················· ···················· ···················· ····················
K 5872 hCoV-19/Indonesia/SA-EIJK-12/2020|EPI_ISL_568694|2 ···················· ···················· ···················· ···················· ···················· ····················
K 5873 hCoV-19/USA/CA-IGI-0239/2020|EPI_ISL_569945|2020-0 ···················· ···················· ···················· ···················· ···················· ····················
K 5874 hCoV-19/USA/CA-IGI-0243/2020|EPI_ISL_569947|2020-0 ···················· ···················· ···················· ···················· ···················· ····················
K 5875 hCoV-19/Canada/ON-UHTC-0174/2020|EPI_ISL_569949|20 ···················· ···················· ···················· ···················· ···················· ····················
K 5876 hCoV-19/Canada/ON-UHTC-0188/2020|EPI_ISL_569954|20 ···················· ···················· ···················· ···················· ···················· ····················
K 5877 hCoV-19/Canada/ON-UHTC-0186/2020|EPI_ISL_569955|20 ···················· ···················· ···················· ···················· ···················· ····················
K 5878 hCoV-19/USA/WA-UW-12911/2020|EPI_ISL_570929|2020-0 ···················· ·A·················· ···················· ···················· ···················· ····················
K 5879 hCoV-19/USA/WA-UW-12921/2020|EPI_ISL_570930|2020-0 ···················· ···················· ···················· ···················· ···················· ····················
K 5880 hCoV-19/USA/WA-UW-12924/2020|EPI_ISL_570931|2020-0 ···················· ···················· ···················· ···················· ···················· ····················
K 5881 hCoV-19/USA/WA-UW-12926/2020|EPI_ISL_570932|2020-0 ···················· ···················· ···················· ···················· ···················· ····················
K 5882 hCoV-19/USA/WA-UW-12930/2020|EPI_ISL_570933|2020-0 ···················· ···················· ···················· ···················· ···················· ····················

| | | | | | | | | |
|---|---|---|---|---|---|---|---|---|
| K | 5883 | hCoV-19/USA/WA-UW-12934/2020\|EPI_ISL_570934\|2020-0 | ···················· | ···················· | ···················· | ···················· | ···················· | ···················· |
| K | 5884 | hCoV-19/USA/WA-UW-12936/2020\|EPI_ISL_570935\|2020-0 | ···················· | ···················· | ···················· | ···················· | ···················· | ···················· |
| K | 5885 | hCoV-19/USA/WA-UW-12948/2020\|EPI_ISL_570936\|2020-0 | ···················· | ···················· | ···················· | ···················· | ···················· | ···················· |
| K | 5886 | hCoV-19/USA/WA-UW-12957/2020\|EPI_ISL_570937\|2020-0 | ···················· | ···················· | ···················· | ···················· | ···················· | ···················· |
| K | 5887 | hCoV-19/USA/WA-UW-12962/2020\|EPI_ISL_570938\|2020-0 | ···················· | ···················· | ···················· | ···················· | ···················· | ···················· |
| K | 5888 | hCoV-19/USA/WA-UW-12978/2020\|EPI_ISL_570939\|2020-0 | ···················· | ···················· | ···················· | ···················· | ···················· | ···················· |
| K | 5889 | hCoV-19/Gambia/NPHL-247/2020\|EPI_ISL_561257\|2020 | ···················· | ···················· | ···················· | ···················· | ···················· | ···················· |
| K | 5890 | hCoV-19/Gambia/NPHL-2473/2020\|EPI_ISL_561260\|2020- | ···················· | ···················· | ···················· | ···················· | ···················· | ···················· |
| K | 5891 | hCoV-19/Gambia/NPHL-2500/2020\|EPI_ISL_561263\|2020- | ···················· | ···················· | ···················· | ···················· | ···················· | ···················· |
| K | 5892 | hCoV-19/Gambia/NPHL-2524/2020\|EPI_ISL_561268\|2020- | ···················· | ···················· | ···················· | ···················· | ···················· | ···················· |
| K | 5893 | hCoV-19/Gambia/NPHL-253/2020\|EPI_ISL_561270\|2020 | ···················· | ···················· | ···················· | ···················· | ···················· | ···················· |
| K | 5894 | hCoV-19/Gambia/NPHL-255/2020\|EPI_ISL_561273\|2020 | ···················· | ···················· | ···················· | ···················· | ···················· | ···················· |
| K | 5895 | hCoV-19/Gambia/NPHL-256/2020\|EPI_ISL_561277\|2020 | ···················· | ···················· | ···················· | ···················· | ···················· | ···················· |
| K | 5896 | hCoV-19/Gambia/NPHL-2567/2020\|EPI_ISL_561279\|2020 | ···················· | ···················· | ···················· | ···················· | ···················· | ···················· |
| K | 5897 | hCoV-19/Gambia/NPHL-2588/2020\|EPI_ISL_561282\|2020 | ···················· | ···················· | ···················· | ···················· | ···················· | ···················· |
| K | 5898 | hCoV-19/Gambia/NPHL-2615/2020\|EPI_ISL_561283\|2020- | ···················· | ···················· | ···················· | ···················· | ···················· | ···················· |
| K | 5899 | hCoV-19/Gambia/NPHL-2626/2020\|EPI_ISL_561284\|2020 | ···················· | ···················· | ···················· | ···················· | ···················· | ···················· |
| K | 5900 | hCoV-19/Gambia/NPHL-2640/2020\|EPI_ISL_561285\|2020- | ···················· | ···················· | ···················· | ···················· | ···················· | ···················· |
| K | 5901 | hCoV-19/Gambia/NPHL-2706/2020\|EPI_ISL_561287\|2020 | ···················· | ···················· | ···················· | ···················· | ···················· | ···················· |
| K | 5902 | hCoV-19/Gambia/NPHL-2712/2020\|EPI_ISL_561288\|2020- | ···················· | ···················· | ···················· | ···················· | ···················· | ···················· |
| K | 5903 | hCoV-19/Gambia/NPHL-2727/2020\|EPI_ISL_561289\|2020- | ···················· | ···················· | ···················· | ···················· | ···················· | ···················· |
| K | 5904 | hCoV-19/Gambia/NPHL-2729/2020\|EPI_ISL_561290\|2020- | ···················· | ···················· | ···················· | ···················· | ···················· | ···················· |
| K | 5905 | hCoV-19/Gambia/NPHL-2736/2020\|EPI_ISL_561291\|2020- | ···················· | ···················· | ···················· | ···················· | ···················· | ···················· |
| K | 5906 | hCoV-19/Gambia/NPHL-2759/2020\|EPI_ISL_561292\|2020- | ···················· | ···················· | ···················· | ···················· | ···················· | ···················· |
| K | 5907 | hCoV-19/Gambia/NPHL-2762/2020\|EPI_ISL_561293\|2020- | ···················· | ···················· | ···················· | ···················· | ···················· | ···················· |
| K | 5908 | hCoV-19/Gambia/NPHL-2767/2020\|EPI_ISL_561294\|2020- | ···················· | ···················· | ···················· | ···················· | ···················· | ···················· |
| K | 5909 | hCoV-19/Gambia/NPHL-2768/2020\|EPI_ISL_561295\|2020- | ···················· | ···················· | ···················· | ···················· | ···················· | ···················· |
| K | 5910 | hCoV-19/Gambia/NPHL-277/2020\|EPI_ISL_561296\|2020 | ···················· | ···················· | ···················· | ···················· | ···················· | ···················· |
| K | 5911 | hCoV-19/Gambia/NPHL-2785/2020\|EPI_ISL_561297\|2020- | ···················· | ···················· | ···················· | ···················· | ···················· | ···················· |
| K | 5912 | hCoV-19/Gambia/NPHL-2790/2020\|EPI_ISL_561298\|2020 | ···················· | ···················· | ···················· | ···················· | ···················· | ···················· |
| K | 5913 | hCoV-19/Gambia/NPHL-2825/2020\|EPI_ISL_561299\|2020 | ···················· | ···················· | ···················· | ···················· | ···················· | ···················· |
| K | 5914 | hCoV-19/Australia/VIC10692/2020\|EPI_ISL_562591\|202 | ···················· | ···················· | ···················· | ···················· | ···················· | ···················· |
| K | 5915 | hCoV-19/Australia/VIC10789/2020\|EPI_ISL_562592\|202 | ···················· | ···················· | ···················· | ···················· | ···················· | ···················· |
| K | 5916 | hCoV-19/Australia/VIC10895/2020\|EPI_ISL_562593\|202 | ···················· | ···················· | ···················· | ···················· | ···················· | ···················· |
| K | 5917 | hCoV-19/Australia/VIC12847/2020\|EPI_ISL_562594\|202 | ···················· | ···················· | ···················· | ···················· | ···················· | ···················· |
| K | 5918 | hCoV-19/Australia/VIC13161/2020\|EPI_ISL_562595\|202 | ···················· | ···················· | ···················· | ···················· | ···················· | ···················· |
| K | 5919 | hCoV-19/Australia/VIC10076/2020\|EPI_ISL_562596\|202 | ···················· | ···················· | ···················· | ···················· | ···················· | ···················· |
| K | 5920 | hCoV-19/Australia/VIC10131/2020\|EPI_ISL_562597\|202 | ···················· | ···················· | ···················· | ···················· | ···················· | ···················· |
| K | 5921 | hCoV-19/Australia/VIC10355/2020\|EPI_ISL_562598\|202 | ···················· | ···················· | ···················· | ···················· | ···················· | ···················· |
| K | 5922 | hCoV-19/Australia/VIC10479/2020\|EPI_ISL_562599\|202 | ···················· | ···················· | ···················· | ···················· | ···················· | ···················· |
| K | 5923 | hCoV-19/Australia/VIC13787/2020\|EPI_ISL_563837\|202 | ···················· | ···················· | ···················· | ···················· | ···················· | ···················· |
| K | 5924 | hCoV-19/Australia/VIC13788/2020\|EPI_ISL_563838\|202 | ···················· | ···················· | ···················· | ···················· | ···················· | ···················· |
| K | 5925 | hCoV-19/Australia/VIC13790/2020\|EPI_ISL_563840\|202 | ···················· | ···················· | ···················· | ···················· | ···················· | ···················· |
| K | 5926 | hCoV-19/Australia/VIC13821/2020\|EPI_ISL_563841\|202 | ···················· | ···················· | ···················· | ···················· | ···················· | ···················· |
| K | 5927 | hCoV-19/Australia/VIC13822/2020\|EPI_ISL_563842\|202 | ···················· | ···················· | ···················· | ···················· | ···················· | ···················· |
| K | 5928 | hCoV-19/Australia/VIC13824/2020\|EPI_ISL_563843\|202 | ···················· | ···················· | ···················· | ···················· | ···················· | ···················· |
| K | 5932 | hCoV-19/Australia/VIC13829/2020\|EPI_ISL_563847\|202 | ···················· | ···················· | ···················· | ···················· | ···················· | ···················· |
| K | 5933 | hCoV-19/Australia/VIC13831/2020\|EPI_ISL_563848\|202 | ···················· | ···················· | ···················· | ···················· | ···················· | ···················· |
| K | 5934 | hCoV-19/Australia/VIC13834/2020\|EPI_ISL_563849\|202 | ···················· | ···················· | ···················· | ···················· | ···················· | ···················· |
| K | 5935 | hCoV-19/Australia/VIC13835/2020\|EPI_ISL_563850\|202 | ···················· | ···················· | ···················· | ···················· | ···················· | ···················· |
| K | 5936 | hCoV-19/USA/MI-MDHHS-SC21968/2020\|EPI_ISL_566019\|2 | ···················· | ···················· | ···················· | ···················· | ···················· | ···················· |
| K | 5937 | hCoV-19/USA/MI-MDHHS-SC21969/2020\|EPI_ISL_566020\|2 | ···················· | ···················· | ···················· | ···················· | ···················· | ···················· |
| K | 5938 | hCoV-19/USA/MI-MDHHS-SC21970/2020\|EPI_ISL_566021\|2 | ···················· | ···················· | ···················· | ···················· | ···················· | ···················· |
| K | 5939 | hCoV-19/USA/MI-MDHHS-SC21971/2020\|EPI_ISL_566022\|2 | ···················· | ·A·················· | ···················· | ···················· | ···················· | ···················· |
| K | 5940 | hCoV-19/USA/MI-MDHHS-SC21972/2020\|EPI_ISL_566023\|2 | ···················· | ···················· | ···················· | ···················· | ···················· | ···················· |
| K | 5941 | hCoV-19/USA/MI-MDHHS-SC21973/2020\|EPI_ISL_566024\|2 | ···················· | ···················· | ···················· | ···················· | ···················· | ···················· |
| K | 5942 | hCoV-19/USA/MI-MDHHS-SC21974/2020\|EPI_ISL_566025\|2 | ···················· | ···················· | ···················· | ···················· | ···················· | ···················· |
| K | 5943 | hCoV-19/USA/MI-MDHHS-SC21975/2020\|EPI_ISL_566026\|2 | ···················· | ···················· | ···················· | ···················· | ···················· | ···················· |
| K | 5944 | hCoV-19/USA/MI-MDHHS-SC21977/2020\|EPI_ISL_566027\|2 | ···················· | ···················· | ···················· | ···················· | ···················· | ···················· |
| K | 5945 | hCoV-19/USA/MI-MDHHS-SC21978/2020\|EPI_ISL_566028\|2 | ···················· | ···················· | ···················· | ···················· | ···················· | ···················· |
| K | 5946 | hCoV-19/England/QEUH-9CA449/2020\|EPI_ISL_567300\|20 | ···················· | ···················· | ···················· | ···················· | ···················· | ···················· |
| K | 5947 | hCoV-19/England/QEUH-9CA106/2020\|EPI_ISL_567301\|20 | ···················· | ·A·················· | ···················· | ···················· | ···················· | ··A················· |
| K | 5948 | hCoV-19/England/QEUH-9C9BFA/2020\|EPI_ISL_567302\|20 | ···················· | ···················· | ···················· | ···················· | ···················· | ···················· |
| K | 5949 | hCoV-19/England/QEUH-9CA810/2020\|EPI_ISL_567303\|20 | ···················· | ···················· | ···················· | ···················· | ···················· | ···················· |
| K | 5950 | hCoV-19/Wales/QEUH-9CA485/2020\|EPI_ISL_567304\|2020 | ···················· | ···················· | ···················· | ···················· | ···················· | ···················· |
| K | 5951 | hCoV-19/England/QEUH-9C93D4/2020\|EPI_ISL_567306\|20 | ···················· | ···················· | ···················· | ···················· | ···················· | ···················· |
| K | 5952 | hCoV-19/England/QEUH-9CA3D3/2020\|EPI_ISL_567307\|20 | ···················· | ···················· | ···················· | ···················· | ···················· | ···················· |
| K | 5953 | hCoV-19/England/QEUH-9C9705/2020\|EPI_ISL_567308\|20 | ···················· | ···················· | ···················· | ···················· | ···················· | ···················· |
| K | 5954 | hCoV-19/England/QEUH-9CA986/2020\|EPI_ISL_567309\|20 | ···················· | ···················· | ···················· | ···················· | ···················· | ···················· |
| K | 5955 | hCoV-19/Wales/QEUH-9C98D5/2020\|EPI_ISL_567310\|2020 | ···················· | ···················· | ···················· | ···················· | ···················· | ···················· |
| K | 5956 | hCoV-19/England/QEUH-9C9B27/2020\|EPI_ISL_567311\|20 | ···················· | ···················· | ···················· | ···················· | ···················· | ···················· |
| K | 5957 | hCoV-19/England/QEUH-9CA29A/2020\|EPI_ISL_567312\|20 | ···················· | ···················· | ···················· | ···················· | ···················· | ···················· |
| K | 5958 | hCoV-19/England/QEUH-9C9CCA/2020\|EPI_ISL_567313\|20 | ···················· | ·A·················· | ···················· | ···················· | ···················· | ··A················· |
| K | 5959 | hCoV-19/England/QEUH-9CA1CA/2020\|EPI_ISL_567314\|20 | ···················· | ···················· | ···················· | ···················· | ···················· | ···················· |
| K | 5960 | hCoV-19/England/QEUH-9CA7AA/2020\|EPI_ISL_567315\|20 | ···················· | ···················· | ···················· | ···················· | ···················· | ···················· |
| K | 5961 | hCoV-19/England/QEUH-9C9FA3/2020\|EPI_ISL_567316\|20 | ···················· | ···················· | ···················· | ···················· | ···················· | ···················· |
| K | 5962 | hCoV-19/England/QEUH-9C9310/2020\|EPI_ISL_567317\|20 | ···················· | ···················· | ···················· | ···················· | ···················· | ···················· |
| K | 5963 | hCoV-19/England/QEUH-9C9FB2/2020\|EPI_ISL_567320\|20 | ···················· | ···················· | ···················· | ···················· | ···················· | ···················· |
| K | 5964 | hCoV-19/England/QEUH-9CA17F/2020\|EPI_ISL_567322\|20 | ···················· | ···················· | ···················· | ···················· | ···················· | ···················· |
| K | 5965 | hCoV-19/England/QEUH-9C9F76/2020\|EPI_ISL_567323\|20 | ···················· | ···················· | ···················· | ···················· | ···················· | ···················· |

```
K    5966 hCoV-19/England/QEUH-9CA625/2020|EPI_ISL_567324|20   ....................  ....................  .....................  ....................  ....................  ....................
K    5967 hCoV-19/England/QEUH-9CA86B/2020|EPI_ISL_567325|20   ....................  ....................  .....................  ....................  ....................  ....................
K    5968 hCoV-19/England/QEUH-9C95CF/2020|EPI_ISL_567327|20   ....................  ....................  .....................  ....................  ....................  ....................
K    5969 hCoV-19/England/QEUH-9C9AB1/2020|EPI_ISL_567329|20   ....................  .A..................  .....................  ....................  ....................  ..A.................
K    5970 hCoV-19/England/QEUH-9C9987/2020|EPI_ISL_567330|20   ....................  ....................  .....................  ....................  ....................  ....................
K    5971 hCoV-19/England/QEUH-9C9A48/2020|EPI_ISL_567331|20   ....................  ....................  .....................  ....................  ....................  ....................
K    5972 hCoV-19/England/QEUH-9C943B/2020|EPI_ISL_567332|20   ....................  ....................  .....................  ....................  ....................  ....................
K    5973 hCoV-19/England/QEUH-9C9C42/2020|EPI_ISL_567333|20   ....................  ....................  .....................  ....................  ....................  ....................
K    5974 hCoV-19/England/QEUH-9C9723/2020|EPI_ISL_567334|20   ....................  ....................  .....................  ....................  ......T.............  ....................
K    5975 hCoV-19/England/QEUH-9CA722/2020|EPI_ISL_567335|20   ....................  ....................  .....................  ....................  ....................  ....................
K    5976 hCoV-19/England/QEUH-9C9495/2020|EPI_ISL_567336|20   ....................  ....................  .....................  ....................  ....................  ....................
K    5977 hCoV-19/England/QEUH-9C9E2E/2020|EPI_ISL_567337|20   ....................  ....................  .....................  ....................  ....................  ....................
K    5978 hCoV-19/England/QEUH-9CA661/2020|EPI_ISL_567338|20   ....................  ....................  .....................  ....................  ....................  ....................
K    5979 hCoV-19/England/QEUH-9C934D/2020|EPI_ISL_567339|20   ....................  ....................  .....................  ....................  ....................  ....................
K    5980 hCoV-19/England/QEUH-9CA582/2020|EPI_ISL_567340|20   ....................  ....................  .....................  ....................  ....................  ....................
K    5981 hCoV-19/England/QEUH-9CA93B/2020|EPI_ISL_567341|20   ....................  ....................  .....................  ....................  ....................  ....................
K    5982 hCoV-19/England/QEUH-9C9C33/2020|EPI_ISL_567345|20   ....................  ....................  .....................  ....................  ....................  ....................
K    5983 hCoV-19/England/QEUH-9C95DE/2020|EPI_ISL_567346|20   ....................  ....................  .....................  ....................  ....................  ....................
K    5984 hCoV-19/England/QEUH-9C9FFE/2020|EPI_ISL_567347|20   ....................  ....................  .....................  ....................  ....................  ....................
K    5985 hCoV-19/England/QEUH-9C9C51/2020|EPI_ISL_567348|20   ....................  ....................  .....................  ....................  ....................  ....................
K    5986 hCoV-19/England/QEUH-9C9468/2020|EPI_ISL_567349|20   ....................  ....................  .....................  ....................  ....................  ....................
K    5987 hCoV-19/England/QEUH-9CA5FB/2020|EPI_ISL_567350|20   ....................  ......C.............  .....................  ....................  ....................  ....................
K    5988 hCoV-19/England/QEUH-9C9B36/2020|EPI_ISL_567352|20   ....................  ....................  .....................  ....................  ....................  ....................
K    5989 hCoV-19/England/QEUH-9CA133/2020|EPI_ISL_567353|20   ....................  ....................  .....................  ....................  ....................  ....................
K    5990 hCoV-19/England/QEUH-9C9F3A/2020|EPI_ISL_567355|20   ....................  .A..................  .....................  ....................  ....................  ....................
K    5991 hCoV-19/England/QEUH-9CA027/2020|EPI_ISL_567356|20   ....................  ....................  .....................  ....................  ....................  ....................
K    5992 hCoV-19/England/QEUH-9C93A7/2020|EPI_ISL_567357|20   ....................  ....................  .....................  ....................  ....................  ....................
K    5993 hCoV-19/England/QEUH-9C9F49/2020|EPI_ISL_567358|20   ....................  ....................  .....................  ....................  ....................  ....................
K    5994 hCoV-19/England/QEUH-9C9CD9/2020|EPI_ISL_567359|20   ....................  ....................  .....................  ....................  ....................  ....................
K    5995 hCoV-19/USA/FL-BPHL-1505/2020|EPI_ISL_568603|2020-   ....................  ....................  .....................  ....................  ....................  ....................
K    5996 hCoV-19/USA/FL-BPHL-1506/2020|EPI_ISL_568604|2020-   ....................  ....................  .....................  ....................  ....................  ....................
K    5997 hCoV-19/USA/FL-BPHL-1508/2020|EPI_ISL_568606|2020-   ....................  .............A......  .....................  ....................  ....................  ....................
K    5998 hCoV-19/USA/FL-BPHL-1509/2020|EPI_ISL_568607|2020-   ....................  ....................  .....................  ....................  ....................  ....................
K    5999 hCoV-19/USA/FL-BPHL-1510/2020|EPI_ISL_568608|2020-   ....................  ....................  .....................  ....................  ....................  ....................
K    6000 hCoV-19/USA/FL-BPHL-1512/2020|EPI_ISL_568610|2020-   ....................  ....................  .....................  ....................  ....................  ....................
K    6001 hCoV-19/USA/FL-BPHL-1514/2020|EPI_ISL_568612|2020-   ....................  ....................  .....................  ....................  ....................  ....................
K    6002 hCoV-19/USA/FL-BPHL-1527/2020|EPI_ISL_568625|2020-   ....................  ....................  .....................  ....................  ....................  ....................
K    6003 hCoV-19/USA/FL-BPHL-1528/2020|EPI_ISL_568626|2020-   ....................  ....................  .....................  ....................  ....................  ....................
K    6004 hCoV-19/USA/FL-BPHL-1529/2020|EPI_ISL_568627|2020-   ....................  ....................  .....................  ....................  ....................  ....................
K    6005 hCoV-19/USA/FL-BPHL-1530/2020|EPI_ISL_568628|2020-   ....................  .A..................  .....................  ....................  ....................  ....................
K    6006 hCoV-19/USA/FL-BPHL-1535/2020|EPI_ISL_568633|2020-   ....................  ....T...............  .....................  ....................  N...................  ....................
K    6007 hCoV-19/USA/FL-BPHL-1539/2020|EPI_ISL_568637|2020-   ....................  ....T...............  .....................  ....................  ....................  ....................
K    6009 hCoV-19/USA/FL-BPHL-1541/2020|EPI_ISL_568639|2020-   ....................  ....................  .........T...........  ....................  ....................  ....................
K    6010 hCoV-19/USA/FL-BPHL-1542/2020|EPI_ISL_568640|2020-   ....................  ....................  .....................  ....................  ....................  ....................
K    6011 hCoV-19/USA/FL-BPHL-1546/2020|EPI_ISL_568644|2020-   ....................  ....................  .....................  ....................  ....................  ....................
K    6012 hCoV-19/USA/FL-BPHL-1547/2020|EPI_ISL_568645|2020-   ....................  ....................  .....................  ....................  NNN.................  ....................            1
K    6013 hCoV-19/USA/FL-BPHL-1550/2020|EPI_ISL_568648|2020-   ....................  ....................  .....................  ....................  ....................  ....................
K    6014 hCoV-19/USA/FL-BPHL-1551/2020|EPI_ISL_568649|2020-   ....................  .............A......  .....................  ....................  ....................  ....................
K    6015 hCoV-19/USA/FL-BPHL-1553/2020|EPI_ISL_568651|2020-   ....................  ....................  .....................  ....................  ....................  ....................
K    6016 hCoV-19/USA/FL-BPHL-1554/2020|EPI_ISL_568652|2020-   ....................  ....................  .....................  ....................  ....................  ....................
K    6017 hCoV-19/USA/FL-BPHL-1556/2020|EPI_ISL_568654|2020-   ....................  ....................  .....................  ....................  ....................  ....................
K    6018 hCoV-19/USA/FL-BPHL-1558/2020|EPI_ISL_568656|2020-   ....................  ....................  .....................  ....................  ....................  ....................
K    6019 hCoV-19/USA/FL-BPHL-1559/2020|EPI_ISL_568657|2020-   ....................  ....................  .....................  ....................  ....................  ....................
K    6020 hCoV-19/USA/FL-BPHL-1560/2020|EPI_ISL_568658|2020-   ....................  ....................  .....................  ....................  ....................  ....................
K    6023 hCoV-19/USA/FL-BPHL-1563/2020|EPI_ISL_568661|2020-   ....................  ....................  .....................  ....................  ....................  ....................
K    6024 hCoV-19/USA/FL-BPHL-1564/2020|EPI_ISL_568662|2020-   ....................  ....................  .....................  ....................  ....................  ....................
K    6026 hCoV-19/USA/FL-BPHL-1566/2020|EPI_ISL_568664|2020-   ....................  ....................  .....................  ....................  ....................  ....................
K    6027 hCoV-19/USA/FL-BPHL-1567/2020|EPI_ISL_568665|2020-   ....................  ....................  .....................  ....................  ....................  ....................
K    6028 hCoV-19/USA/FL-BPHL-1568/2020|EPI_ISL_568666|2020-   ....................  ....................  .....................  ....................  ....................  ....................
K    6029 hCoV-19/USA/FL-BPHL-1569/2020|EPI_ISL_568667|2020-   ....................  .............A......  .....................  ....................  ....................  ....................
K    6030 hCoV-19/USA/FL-BPHL-1570/2020|EPI_ISL_568668|2020-   ....................  ....................  .....................  ....................  ....................  ....................
K    6031 hCoV-19/USA/FL-BPHL-1571/2020|EPI_ISL_568669|2020-   ....................  ....................  .....................  ....................  ....................  ....................
K    6032 hCoV-19/USA/FL-BPHL-1573/2020|EPI_ISL_568671|2020-   ....................  ....................  .....................  ....................  ....................  ....................
K    6034 hCoV-19/USA/FL-BPHL-1578/2020|EPI_ISL_568676|2020-   ....................  ....................  .....................  ....................  ....................  ....................
K    6035 hCoV-19/USA/FL-BPHL-1579/2020|EPI_ISL_568677|2020-   ....................  .............A......  .....................  ....................  ....................  ....................
K    6036 hCoV-19/USA/FL-BPHL-1580/2020|EPI_ISL_568678|2020-   ....................  ....................  .....................  ....................  ....................  ....................
K    6037 hCoV-19/USA/FL-BPHL-1581/2020|EPI_ISL_568679|2020-   ....................  ....................  .....................  ....................  ....................  ....................
K    6038 hCoV-19/USA/FL-BPHL-1583/2020|EPI_ISL_568681|2020-   ....................  ....................  .....................  ....................  ....................  ....................
K    6039 hCoV-19/Indonesia/JI-EIJK-10/2020|EPI_ISL_568686|2   ....................  ....................  .....................  ....................  ....................  ....................
K    6040 hCoV-19/Indonesia/JK-EIJK-23/2020|EPI_ISL_568687|2   ....................  ....................  .....................  ....................  ....................  ....................
K    6041 hCoV-19/Indonesia/JK-EIJK-32/2020|EPI_ISL_568688|2   ....................  ....................  .....................  ....................  ....................  ....................
K    6042 hCoV-19/Indonesia/JK-EIJK-38/2020|EPI_ISL_568689|2   ....................  ....................  .....................  ....................  ....................  ....................
K    6043 hCoV-19/Kenya/C571/2020|EPI_ISL_568702|2020-04-12    ....................  ....................  .....................  ....................  ....................  ....................
K    6044 hCoV-19/USA/CA-IGI-0207/2020|EPI_ISL_569934|2020-0   ....................  ....................  .....................  ....................  ....................  ....................
K    6045 hCoV-19/USA/CA-IGI-0208/2020|EPI_ISL_569935|2020-0   ....................  ....................  .....................  ....................  ....................  ....................
K    6046 hCoV-19/USA/CA-IGI-0219/2020|EPI_ISL_569939|2020-0   ....................  ....................  .....................  ....................  ....................  ....................
K    6047 hCoV-19/USA/CA-IGI-0221/2020|EPI_ISL_569940|2020-0   ....................  .A..................  .....................  ....................  ....................  ....................
K    6048 hCoV-19/USA/CA-IGI-0223/2020|EPI_ISL_569941|2020-0   ....................  ....................  .....................  ....................  ....................  ....................
K    6049 hCoV-19/USA/CA-IGI-0226/2020|EPI_ISL_569942|2020-0   ....................  ....................  .....................  ....................  ....................  ....................
K    6050 hCoV-19/USA/CA-IGI-0227/2020|EPI_ISL_569943|2020-0   ....................  ....................  .....................  ....................  ....................  ....................
```

| | | | | | | | | |
|---|---|---|---|---|---|---|---|---|
| K | 6051 | hCoV-19/Canada/ON-UHTC-0194/2020\|EPI_ISL_569958\|20 | ···················· | ···················· | ···················· | ···················· | ···················· | ···················· |
| K | 6052 | hCoV-19/Canada/ON-UHTC-0200/2020\|EPI_ISL_569961\|20 | ···················· | ···················· | ···················· | ···················· | ···················· | ···················· |
| K | 6053 | hCoV-19/Canada/ON-UHTC-0189/2020\|EPI_ISL_569965\|20 | ···················· | ···················· | ···················· | ···················· | ···················· | ···················· |
| K | 6054 | hCoV-19/Canada/ON-UHTC-0180/2020\|EPI_ISL_569966\|20 | ···················· | ···················· | ···················· | ···················· | ···················· | ···················· |
| K | 6055 | hCoV-19/Canada/ON-UHTC-0177/2020\|EPI_ISL_569968\|20 | ···················· | ···················· | ···················· | ···················· | ···················· | ···················· |
| K | 6056 | hCoV-19/Canada/ON-UHTC-0199/2020\|EPI_ISL_569969\|20 | ···················· | ·A·················· | ···················· | ···················· | ···················· | ··A················· |
| K | 6057 | hCoV-19/Canada/ON-UHTC-0176/2020\|EPI_ISL_569970\|20 | ···················· | ···················· | ···················· | ···················· | ···················· | ···················· |
| K | 6058 | hCoV-19/Canada/ON-UHTC-0178/2020\|EPI_ISL_569971\|20 | ···················· | ···················· | ···················· | ···················· | ···················· | ···················· |
| K | 6059 | hCoV-19/Canada/ON-UHTC-0202/2020\|EPI_ISL_569972\|20 | ···················· | ···················· | ···················· | ···················· | ···················· | ···················· |
| K | 6060 | hCoV-19/Canada/ON-UHTC-0175/2020\|EPI_ISL_569973\|20 | ···················· | ···················· | ···················· | ···················· | ···················· | ···················· |
| K | 6061 | hCoV-19/Canada/ON-UHTC-0203/2020\|EPI_ISL_569975\|20 | ···················· | ···················· | ···················· | ···················· | ···················· | ···················· |
| K | 6062 | hCoV-19/Canada/ON-UHTC-0179/2020\|EPI_ISL_569976\|20 | ···················· | ···················· | ···················· | ···················· | ···················· | ···················· |
| K | 6063 | hCoV-19/Canada/ON-UHTC-0197/2020\|EPI_ISL_569978\|20 | ···················· | ···················· | ···················· | ···················· | ···················· | ···················· |
| K | 6064 | hCoV-19/Canada/ON-UHTC-0293/2020\|EPI_ISL_569980\|20 | ···················· | ···················· | ···················· | ···················· | ···················· | ···················· |
| K | 6065 | hCoV-19/Canada/ON-UHTC-0283/2020\|EPI_ISL_569981\|20 | ···················· | ···················· | ···················· | ···················· | ···················· | ···················· |
| K | 6066 | hCoV-19/Canada/ON-UHTC-0285/2020\|EPI_ISL_569982\|20 | ···················· | ···················· | ···················· | ···················· | ···················· | ···················· |
| K | 6067 | hCoV-19/Canada/ON-UHTC-0284/2020\|EPI_ISL_569983\|20 | ···················· | ···················· | ···················· | ···················· | ···················· | ···················· |
| K | 6068 | hCoV-19/Canada/ON-UHTC-0296/2020\|EPI_ISL_569986\|20 | ···················· | ···················· | ···················· | ···················· | ···················· | ···················· |
| K | 6069 | hCoV-19/Canada/ON-UHTC-0286/2020\|EPI_ISL_569987\|20 | ···················· | ···················· | ···················· | ···················· | ···················· | ···················· |
| K | 6070 | hCoV-19/Canada/ON-UHTC-0291/2020\|EPI_ISL_569991\|20 | ···················· | ···················· | ···················· | ···················· | ···················· | ···················· |
| K | 6071 | hCoV-19/Canada/ON-UHTC-0282/2020\|EPI_ISL_569992\|20 | ···················· | ···················· | ···················· | ···················· | ···················· | ···················· |
| K | 6072 | hCoV-19/Canada/ON-UHTC-0288/2020\|EPI_ISL_569993\|20 | ···················· | ···················· | ···················· | ···················· | ···················· | ···················· |
| K | 6073 | hCoV-19/Canada/ON-UHTC-0258/2020\|EPI_ISL_569995\|20 | ···················· | ···················· | ···················· | ···················· | ···················· | ···················· |
| K | 6074 | hCoV-19/Canada/ON-UHTC-0267/2020\|EPI_ISL_569996\|20 | ···················· | ···················· | ···················· | ···················· | ···················· | ···················· |
| K | 6075 | hCoV-19/Canada/ON-UHTC-0272/2020\|EPI_ISL_569998\|20 | ···················· | ···················· | ···················· | ···················· | ···················· | ···················· |
| K | 6076 | hCoV-19/USA/WA-UW-12879/2020\|EPI_ISL_570920\|2020-0 | ···················· | ···················· | ···················· | ···················· | ···················· | ···················· |
| K | 6077 | hCoV-19/USA/WA-UW-12881/2020\|EPI_ISL_570921\|2020-0 | ···················· | ···················· | ···················· | ···················· | ···················· | ···················· |
| K | 6078 | hCoV-19/USA/WA-UW-12883/2020\|EPI_ISL_570922\|2020-0 | ···················· | ···················· | ···················· | ···················· | ···················· | ···················· |
| K | 6079 | hCoV-19/USA/WA-UW-12886/2020\|EPI_ISL_570923\|2020-0 | ···················· | ···················· | ···················· | ···················· | ···················· | ···················· |
| K | 6080 | hCoV-19/USA/WA-UW-12898/2020\|EPI_ISL_570924\|2020-0 | ···················· | ···················· | ···················· | ···················· | ···················· | ···················· |
| K | 6081 | hCoV-19/USA/WA-UW-12901/2020\|EPI_ISL_570925\|2020-0 | ···················· | ···················· | ···················· | ···················· | ···················· | ···················· |
| K | 6082 | hCoV-19/USA/WA-UW-12907/2020\|EPI_ISL_570926\|2020-0 | ···················· | ···················· | ···················· | ···················· | ···················· | ···················· |
| K | 6083 | hCoV-19/USA/WA-UW-12908/2020\|EPI_ISL_570927\|2020-0 | ···················· | ···················· | ···················· | ···················· | ···················· | ···················· |
| K | 6085 | hCoV-19/USA/WA-UW-13003/2020\|EPI_ISL_570940\|2020-0 | ···················· | ···················· | ···················· | ···················· | ···················· | ···················· |
| K | 6086 | hCoV-19/USA/WA-UW-13015/2020\|EPI_ISL_570941\|2020-0 | ···················· | ···················· | ···················· | ···················· | ···················· | ···················· |
| K | 6087 | hCoV-19/USA/WA-UW-13018/2020\|EPI_ISL_570942\|2020-0 | ···················· | ···················· | ···················· | ···················· | ···················· | ···················· |
| K | 6088 | hCoV-19/USA/WA-UW-13027/2020\|EPI_ISL_570943\|2020-0 | ···················· | ···················· | ···················· | ···················· | ···················· | ···················· |
| K | 6089 | hCoV-19/USA/WA-UW-13029/2020\|EPI_ISL_570944\|2020-0 | ···················· | ···················· | ···················· | ···················· | ···················· | ···················· |
| K | 6090 | hCoV-19/USA/WA-UW-13068/2020\|EPI_ISL_570945\|2020-0 | ···················· | ·A·················· | ···················· | ···················· | ···················· | ···················· |
| K | 6091 | hCoV-19/USA/WA-UW-13070/2020\|EPI_ISL_570946\|2020-0 | ···················· | ···················· | ···················· | ···················· | ···················· | ···················· |
| K | 6092 | hCoV-19/USA/WA-UW-13071/2020\|EPI_ISL_570947\|2020-0 | ···················· | ···················· | ···················· | ···················· | ···················· | ···················· |
| K | 6093 | hCoV-19/USA/WA-UW-13080/2020\|EPI_ISL_570948\|2020-0 | ···················· | ·A·················· | ···················· | ···················· | ···················· | ···················· |
| K | 6094 | hCoV-19/USA/WA-UW-13082/2020\|EPI_ISL_570949\|2020-0 | ···················· | ·A·················· | ···················· | ···················· | ···················· | ···················· |
| K | 6095 | hCoV-19/USA/WA-UW-13083/2020\|EPI_ISL_570950\|2020-0 | ···················· | ···················· | ···················· | ···················· | ···················· | ···················· |
| K | 6096 | hCoV-19/USA/WA-UW-13084/2020\|EPI_ISL_570951\|2020-0 | ···················· | ·A·················· | ···················· | ···················· | ···················· | ···················· |
| K | 6097 | hCoV-19/USA/WA-UW-13085/2020\|EPI_ISL_570952\|2020-0 | ···················· | ···················· | ···················· | ···················· | ···················· | ···················· |
| K | 6098 | hCoV-19/USA/WA-UW-13088/2020\|EPI_ISL_570953\|2020-0 | ···················· | ···················· | ···················· | ···················· | ···················· | ···················· |
| K | 6099 | hCoV-19/USA/WA-UW-13104/2020\|EPI_ISL_570954\|2020-0 | ···········T········ | ···················· | ···················· | ···················· | ···················· | ···················· |
| K | 6100 | hCoV-19/USA/WA-UW-13110/2020\|EPI_ISL_570955\|2020-0 | ···················· | ·A·················· | ···················· | ···················· | ···················· | ···················· |
| K | 6101 | hCoV-19/USA/WA-UW-13196/2020\|EPI_ISL_570956\|2020-0 | ···················· | ···················· | ···················· | ···················· | ···················· | ···················· |
| K | 6102 | hCoV-19/USA/WA-UW-13201/2020\|EPI_ISL_570957\|2020-0 | ···················· | ···················· | ···················· | ···················· | ···················· | ···················· |
| K | 6103 | hCoV-19/USA/WA-UW-13207/2020\|EPI_ISL_570958\|2020-0 | ···················· | ·A·················· | ···················· | ···················· | ···················· | ···················· |
| K | 6104 | hCoV-19/USA/WA-UW-13219/2020\|EPI_ISL_570959\|2020-0 | ···················· | ·A·················· | ···················· | ···················· | ···················· | ···················· |
| K | 6105 | hCoV-19/USA/WA-UW-13222/2020\|EPI_ISL_570960\|2020-0 | ···················· | ···················· | ···················· | ···················· | ···················· | ···················· |
| K | 6106 | hCoV-19/USA/WA-UW-13234/2020\|EPI_ISL_570961\|2020-0 | ···················· | ···················· | ···················· | ···················· | ···················· | ···················· |
| K | 6107 | hCoV-19/USA/WA-UW-13249/2020\|EPI_ISL_570962\|2020-0 | ···················· | ···················· | ···················· | ···················· | ···················· | ···················· |
| K | 6108 | hCoV-19/USA/WA-UW-13299/2020\|EPI_ISL_570963\|2020-0 | ···················· | ···················· | ···················· | ···················· | ···················· | ···················· |
| K | 6109 | hCoV-19/USA/WA-UW-13311/2020\|EPI_ISL_570964\|2020-0 | ···················· | ···················· | ···················· | ···················· | ···················· | ···················· |
| K | 6110 | hCoV-19/USA/WA-UW-13319/2020\|EPI_ISL_570965\|2020-0 | ···················· | ···················· | ···················· | ···················· | ···················· | ···················· |
| K | 6111 | hCoV-19/USA/WA-UW-13368/2020\|EPI_ISL_570966\|2020-0 | ···················· | ···················· | ···················· | ···················· | ···················· | ···················· |
| K | 6112 | hCoV-19/USA/WA-UW-13370/2020\|EPI_ISL_570967\|2020-0 | ···················· | ·A·················· | ···················· | ···················· | ···················· | ···················· |
| K | 6113 | hCoV-19/USA/WA-UW-13381/2020\|EPI_ISL_570968\|2020-0 | ···················· | ···················· | ···················· | ···················· | ···················· | ···················· |
| K | 6114 | hCoV-19/USA/WA-UW-13397/2020\|EPI_ISL_570969\|2020-0 | ···················· | ···················· | ···················· | ···················· | ···················· | ···················· |
| K | 6115 | hCoV-19/USA/WA-UW-1766/2020\|EPI_ISL_570970\|2020-04 | ···················· | ···················· | ···················· | ···················· | ···················· | ···················· |
| K | 6116 | hCoV-19/USA/WA-UW-1806/2020\|EPI_ISL_570971\|2020-03 | ···················· | ···················· | ···················· | ···················· | ···················· | ···················· |
| K | 6117 | hCoV-19/USA/WA-UW-1812/2020\|EPI_ISL_570972\|2020-03 | ···················· | ···················· | ···················· | ···················· | ···················· | ···················· |
| K | 6118 | hCoV-19/USA/WA-UW-182/2020\|EPI_ISL_570973\|2020-03- | ···················· | ···················· | ···················· | ···················· | ···················· | ···················· |
| K | 6119 | hCoV-19/USA/WA-UW-185/2020\|EPI_ISL_570974\|2020-03- | ···················· | ···················· | ···················· | ···················· | ···················· | ···················· |
| K | 6120 | hCoV-19/USA/WA-UW-1859/2020\|EPI_ISL_570975\|2020-03 | ···················· | ···················· | ···················· | ···················· | ···················· | ···················· |
| K | 6121 | hCoV-19/USA/WA-UW-191/2020\|EPI_ISL_570976\|2020-03- | ···················· | ···················· | ···················· | ···················· | ···················· | ···················· |
| K | 6122 | hCoV-19/USA/WA-UW-192/2020\|EPI_ISL_570977\|2020-03- | ···················· | ···················· | ···················· | ···················· | ···················· | ···················· |
| K | 6123 | hCoV-19/USA/WA-UW-19384/2020\|EPI_ISL_570978\|2020-0 | ···················· | ···················· | ···················· | ···················· | ···················· | ···················· |
| K | 6124 | hCoV-19/USA/WA-UW-194/2020\|EPI_ISL_570979\|2020-03- | ···················· | ···················· | ···················· | ···················· | ···················· | ···················· |
| K | 6125 | hCoV-19/USA/WA-UW-19418/2020\|EPI_ISL_570980\|2020-0 | ···················· | ···················· | ···················· | ···················· | ···················· | ···················· |
| K | 6126 | hCoV-19/USA/WA-UW-19444/2020\|EPI_ISL_570981\|2020-0 | ···················· | ···················· | ···················· | ···················· | ···················· | ···················· |
| K | 6127 | hCoV-19/USA/WA-UW-19488/2020\|EPI_ISL_570982\|2020-0 | ···················· | ·A·················· | ···················· | ···················· | ···················· | ···················· |
| K | 6128 | hCoV-19/USA/WA-UW-195/2020\|EPI_ISL_570983\|2020-03- | ···················· | ···················· | ···················· | ···················· | ···················· | ···················· |
| K | 6129 | hCoV-19/USA/WA-UW-19676/2020\|EPI_ISL_570984\|2020-0 | ···················· | ···················· | ···················· | ···················· | ···················· | ···················· |
| K | 6130 | hCoV-19/USA/WA-UW-19695/2020\|EPI_ISL_570985\|2020-0 | ···················· | ·A·················· | ···················· | ···················· | ···················· | ···················· |
| K | 6131 | hCoV-19/USA/WA-UW-1997/2020\|EPI_ISL_570986\|2020-03 | ···················· | ···················· | ···················· | ···················· | ···················· | ···················· |

| | | | | | | | | |
|---|---|---|---|---|---|---|---|---|
| K | 6132 | hCoV-19/USA/WA-UW-201/2020\|EPI_ISL_570987\|2020-03- | ···················· | ···················· | ···················· | ···················· | ···················· | ···················· |
| K | 6133 | hCoV-19/USA/WA-UW-21562/2020\|EPI_ISL_570988\|2020-0 | ···················· | ·A·················· | ···················· | ···················· | ···················· | ···················· |
| K | 6134 | hCoV-19/USA/WA-UW-21600/2020\|EPI_ISL_570989\|2020-0 | ···················· | ···················· | ···················· | ···················· | ···················· | ···················· |
| K | 6135 | hCoV-19/USA/WA-UW-21707/2020\|EPI_ISL_570990\|2020-0 | ···················· | ···················· | ···················· | ···················· | ···················· | ···················· |
| K | 6136 | hCoV-19/USA/WA-UW-21710/2020\|EPI_ISL_570991\|2020-0 | ···················· | ···················· | ···················· | ···················· | ···················· | ···················· |
| K | 6137 | hCoV-19/USA/WA-UW-21720/2020\|EPI_ISL_570992\|2020-0 | ···················· | ···················· | ···················· | ···················· | ···················· | ···················· |
| K | 6138 | hCoV-19/USA/WA-UW-21723/2020\|EPI_ISL_570993\|2020-0 | ···················· | ···················· | ···················· | ···················· | ···················· | ···················· |
| K | 6139 | hCoV-19/USA/WA-UW-21727/2020\|EPI_ISL_570994\|2020-0 | ···················· | ···················· | ···················· | ···················· | ···················· | ···················· |
| K | 6140 | hCoV-19/USA/WA-UW-21732/2020\|EPI_ISL_570995\|2020-0 | ···················· | ···················· | ···················· | ···················· | ···················· | ···················· |
| K | 6141 | hCoV-19/USA/WA-UW-21734/2020\|EPI_ISL_570996\|2020-0 | ···················· | ·A·················· | ···················· | ···················· | ···················· | ···················· |
| K | 6142 | hCoV-19/USA/WA-UW-21735/2020\|EPI_ISL_570997\|2020-0 | ···················· | ·A·················· | ···················· | ···················· | ···················· | ···················· |
| K | 6143 | hCoV-19/USA/WA-UW-21736/2020\|EPI_ISL_570998\|2020-0 | ···················· | ···················· | ···················· | ···················· | ···················· | ···················· |
| K | 6144 | hCoV-19/USA/WA-UW-21742/2020\|EPI_ISL_570999\|2020-0 | ···················· | ·A·················· | ···················· | ···················· | ···················· | ···················· |
| K | 6145 | hCoV-19/France/BRE-8932/2020\|EPI_ISL_560600\|2020-0 | ···················· | ···················· | ···················· | ···················· | ···················· | ···················· |
| K | 6146 | hCoV-19/France/BRE-8933/2020\|EPI_ISL_560601\|2020-0 | ···················· | ···················· | ···················· | ···················· | ···················· | ···················· |
| K | 6147 | hCoV-19/France/BRE-8934/2020\|EPI_ISL_560602\|2020-0 | ···················· | ···················· | ···················· | ···················· | ···················· | ···················· |
| K | 6148 | hCoV-19/France/BRE-8935/2020\|EPI_ISL_560603\|2020-0 | ···················· | ···················· | ···················· | ···················· | ···················· | ···················· |
| K | 6149 | hCoV-19/France/BRE-8936/2020\|EPI_ISL_560604\|2020-0 | ···················· | ···················· | ···················· | ···················· | ···················· | ···················· |
| K | 6150 | hCoV-19/France/BRE-8937/2020\|EPI_ISL_560605\|2020-0 | ···················· | ···················· | ···················· | ···················· | ···················· | ···················· |
| K | 6151 | hCoV-19/France/BRE-8938/2020\|EPI_ISL_560606\|2020-0 | ···················· | ···················· | ···················· | ···················· | ···················· | ···················· |
| K | 6152 | hCoV-19/France/BRE-8939/2020\|EPI_ISL_560607\|2020-0 | ···················· | ···················· | ···················· | ···················· | ···················· | ···················· |
| K | 6153 | hCoV-19/France/BRE-8940/2020\|EPI_ISL_560608\|2020-0 | ···················· | ···················· | ···················· | ···················· | ···················· | ···················· |
| K | 6154 | hCoV-19/France/BRE-8941/2020\|EPI_ISL_560609\|2020-0 | ···················· | ·A·················· | ···················· | ···················· | ···················· | ···················· |
| K | 6155 | hCoV-19/France/BRE-8942/2020\|EPI_ISL_560610\|2020-0 | ···················· | ···················· | ···················· | ···················· | ···················· | ···················· |
| K | 6156 | hCoV-19/France/BRE-8943/2020\|EPI_ISL_560611\|2020-0 | ···················· | ···················· | ···················· | ···················· | ···················· | ···················· |
| K | 6157 | hCoV-19/France/BRE-8944/2020\|EPI_ISL_560612\|2020-0 | ···················· | ·A·················· | ···················· | ···················· | ···················· | ···················· |
| K | 6158 | hCoV-19/France/BRE-8946/2020\|EPI_ISL_560613\|2020-0 | ···················· | ···················· | ···················· | ···················· | ···················· | ···················· |
| K | 6159 | hCoV-19/France/BRE-8947/2020\|EPI_ISL_560614\|2020-0 | ···················· | ···················· | ···················· | ···················· | ···················· | ···················· |
| K | 6160 | hCoV-19/France/BRE-8948/2020\|EPI_ISL_560615\|2020-0 | ···················· | ···················· | ···················· | ···················· | ···················· | ···················· |
| K | 6161 | hCoV-19/France/BRE-8949/2020\|EPI_ISL_560616\|2020-0 | ···················· | ···················· | ···················· | ···················· | ···················· | ···················· |
| K | 6162 | hCoV-19/France/BRE-8950/2020\|EPI_ISL_560617\|2020-0 | ···················· | ·A·················· | ···················· | ···················· | ···················· | ···················· |
| K | 6163 | hCoV-19/France/BRE-8951/2020\|EPI_ISL_560618\|2020-0 | ···················· | ···················· | ···················· | ···················· | ···················· | ···················· |
| K | 6164 | hCoV-19/France/GES-8767/2020\|EPI_ISL_560620\|2020-0 | ···················· | ···················· | ···················· | ···················· | ···················· | ···················· |
| K | 6165 | hCoV-19/France/GES-8771/2020\|EPI_ISL_560621\|2020-0 | ···················· | ···················· | ···················· | ···················· | ···················· | ···················· |
| K | 6166 | hCoV-19/France/GES-8774/2020\|EPI_ISL_560622\|2020-0 | ···················· | ···················· | ···················· | ···················· | ···················· | ···················· |
| K | 6167 | hCoV-19/France/GES-8777/2020\|EPI_ISL_560623\|2020-0 | ···················· | ···················· | ···················· | ···················· | ···················· | ···················· |
| K | 6168 | hCoV-19/France/GES-8782/2020\|EPI_ISL_560624\|2020-0 | ···················· | ···················· | ···················· | ···················· | ···················· | ···················· |
| K | 6169 | hCoV-19/France/GES-8783/2020\|EPI_ISL_560625\|2020-0 | ···················· | ···················· | ···················· | ···················· | ···················· | ···················· |
| K | 6170 | hCoV-19/France/GES-8786/2020\|EPI_ISL_560626\|2020-0 | ···················· | ···················· | ···················· | ···················· | ···················· | ···················· |
| K | 6171 | hCoV-19/France/GES-8789/2020\|EPI_ISL_560627\|2020-0 | ···················· | ···················· | ···················· | ···················· | ···················· | ···············A···· |
| K | 6172 | hCoV-19/France/GES-8795/2020\|EPI_ISL_560628\|2020-0 | ···················· | ···················· | ···················· | ···················· | ···················· | ···················· |
| K | 6173 | hCoV-19/France/GES-8799/2020\|EPI_ISL_560629\|2020-0 | ···················· | ···················· | ···················· | ···················· | ···················· | ···················· |
| K | 6174 | hCoV-19/France/GES-8802/2020\|EPI_ISL_560630\|2020-0 | ···················· | ···················· | ···················· | ···················· | ···················· | ···················· |
| K | 6175 | hCoV-19/France/GES-8804/2020\|EPI_ISL_560631\|2020-0 | ···················· | ···················· | ···················· | ···················· | ···················· | ···················· |
| K | 6176 | hCoV-19/France/GES-8805/2020\|EPI_ISL_560632\|2020-0 | ···················· | ···················· | ···················· | ···················· | ···················· | ···················· |
| K | 6177 | hCoV-19/France/GES-8807/2020\|EPI_ISL_560633\|2020-0 | ···················· | ···················· | ···················· | ···················· | ···················· | ···················· |
| K | 6178 | hCoV-19/France/GES-8809/2020\|EPI_ISL_560634\|2020-0 | ···················· | ···················· | ···················· | ···················· | ···················· | ···················· |
| K | 6179 | hCoV-19/France/GES-8815/2020\|EPI_ISL_560635\|2020-0 | ···················· | ···················· | ···················· | ···················· | ···················· | ···················· |
| K | 6180 | hCoV-19/France/IDF-8589/2020\|EPI_ISL_560636\|2020-0 | ···················· | ···················· | ···················· | ···················· | ···················· | ···················· |
| K | 6181 | hCoV-19/France/IDF-8679/2020\|EPI_ISL_560637\|2020-0 | ···················· | ···················· | ···················· | ···················· | ···················· | ···················· |
| K | 6182 | hCoV-19/France/IDF-8683/2020\|EPI_ISL_560638\|2020-0 | ···················· | ···················· | ···············C···· | ···················· | ···················· | ···················· |
| K | 6183 | hCoV-19/France/IDF-8697/2020\|EPI_ISL_560639\|2020-0 | ···················· | ···················· | ···············C···· | ···················· | ···················· | ···················· |
| K | 6184 | hCoV-19/France/IDF-8702/2020\|EPI_ISL_560640\|2020-0 | ···················· | ···················· | ···················· | ···················· | ···················· | ···················· |
| K | 6185 | hCoV-19/France/IDF-8706/2020\|EPI_ISL_560641\|2020-0 | ···················· | ···················· | ···············C···· | ···················· | ···················· | ···················· |
| K | 6186 | hCoV-19/France/IDF-8707/2020\|EPI_ISL_560642\|2020-0 | ···················· | ···················· | ···············C···· | ···················· | ···················· | ···················· |
| K | 6187 | hCoV-19/France/IDF-8761/2020\|EPI_ISL_560643\|2020-0 | ···················· | ···················· | ···················· | ···················· | ···················· | ···················· |
| K | 6188 | hCoV-19/France/IDF-8763/2020\|EPI_ISL_560644\|2020-0 | ···················· | ···················· | ···················· | ···················· | ···················· | ···················· |
| K | 6189 | hCoV-19/France/IDF-8844/2020\|EPI_ISL_560645\|2020-0 | ········T··········· | ···················· | ···················· | ···················· | ···················· | ···················· |
| K | 6190 | hCoV-19/France/IDF-8845/2020\|EPI_ISL_560646\|2020-0 | ········T··········· | ···················· | ···················· | ···················· | ···················· | ···················· |
| K | 6191 | hCoV-19/USA/DE-DHSS-F918664/2020\|EPI_ISL_560647\|20 | ···················· | ···················· | ···················· | ···················· | ···················· | ···················· |
| K | 6192 | hCoV-19/England/GSTT-4106/2020\|EPI_ISL_560650\|2020 | ···················· | ···················· | ···················· | ···················· | ···················· | ···················· |
| K | 6193 | hCoV-19/England/GSTT-5152/2020\|EPI_ISL_560651\|2020 | ···················· | ···················· | ···················· | ···················· | ···················· | ···················· |
| K | 6194 | hCoV-19/England/GSTT-5155/2020\|EPI_ISL_560652\|2020 | ···················· | ···················· | ···················· | ···················· | ···················· | ···················· |
| K | 6195 | hCoV-19/England/GSTT-5179/2020\|EPI_ISL_560656\|2020 | ···················· | ···················· | ···················· | ···················· | ···················· | ···················· |
| K | 6196 | hCoV-19/England/GSTT-5189/2020\|EPI_ISL_560658\|2020 | ···················· | ···················· | ···················· | ···················· | ···················· | ···················· |
| K | 6197 | hCoV-19/England/GSTT-5196/2020\|EPI_ISL_560660\|2020 | ···················· | ···················· | ···················· | ···················· | ···················· | ···················· |
| K | 6198 | hCoV-19/England/GSTT-5203/2020\|EPI_ISL_560661\|2020 | ···················· | ···················· | ···················· | ···················· | ···················· | ···················· |
| K | 6199 | hCoV-19/England/GSTT-5269/2020\|EPI_ISL_560668\|2020 | ···················· | ···················· | ···················· | ···················· | ···················· | ···················· |
| K | 6200 | hCoV-19/England/GSTT-5326/2020\|EPI_ISL_560671\|2020 | ···················· | ···················· | ···················· | ···················· | ···················· | ···················· |
| K | 6201 | hCoV-19/England/GSTT-5340/2020\|EPI_ISL_560672\|2020 | ···················· | ···················· | ···················· | ···················· | ···················· | ···················· |
| K | 6202 | hCoV-19/England/GSTT-5346/2020\|EPI_ISL_560673\|2020 | ···················· | ···················· | ···················· | ···················· | ···················· | ···················· |
| K | 6203 | hCoV-19/England/GSTT-5371/2020\|EPI_ISL_560676\|2020 | ···················· | ···················· | ···················· | ···················· | ···················· | ···················· |
| K | 6204 | hCoV-19/England/GSTT-5393/2020\|EPI_ISL_560678\|2020 | ···················· | ···················· | ···················· | ···················· | ···················· | ···················· |
| K | 6205 | hCoV-19/England/GSTT-5396/2020\|EPI_ISL_560679\|2020 | ···················· | ···················· | ···················· | ···················· | ···················· | ···················· |
| K | 6206 | hCoV-19/Gambia/GC19-3630/2020\|EPI_ISL_561200\|2020- | ···················· | ···················· | ···················· | ···················· | ···················· | ···················· |
| K | 6207 | hCoV-19/Gambia/GC19-3656/2020\|EPI_ISL_561201\|2020- | ···················· | ···················· | ···················· | ···················· | ···················· | ···················· |
| K | 6208 | hCoV-19/Gambia/GC19-3659/2020\|EPI_ISL_561202\|2020- | ···················· | ···················· | ···················· | ···················· | ···················· | ···················· |
| K | 6210 | hCoV-19/Gambia/GC19-3666/2020\|EPI_ISL_561204\|2020- | ···················· | ···················· | ···················· | ···················· | ···················· | ···················· |
| K | 6211 | hCoV-19/Gambia/NPHL-038/2020\|EPI_ISL_561205\|2020 | ···················· | ···················· | ···················· | ···················· | ···················· | ···················· |
| K | 6212 | hCoV-19/Gambia/NPHL-061/2020\|EPI_ISL_561206\|2020 | ···················· | ···················· | ···················· | ···················· | ···················· | ···················· |

| | | | | | | | | | |
|---|---|---|---|---|---|---|---|---|---|
| K | 6213 | hCoV-19/Gambia/NPHL-1168/2020\|EPI_ISL_561212\|2020- | .................... | .................... | .................... | .................... | .................... | .................... | |
| K | 6215 | hCoV-19/Gambia/NPHL-1212/2020\|EPI_ISL_561215\|2020- | .................... | .................... | .................... | .................... | .................... | .................... | |
| K | 6218 | hCoV-19/Gambia/NPHL-1258/2020\|EPI_ISL_561223\|2020- | .................... | .................... | .................... | .................... | .................... | .................... | |
| K | 6219 | hCoV-19/Gambia/NPHL-1424/2020\|EPI_ISL_561229\|2020- | .................... | .................... | .................... | .................... | .................... | .................... | |
| K | 6221 | hCoV-19/Gambia/NPHL-1514/2020\|EPI_ISL_561232\|2020 | .................... | .................... | .................... | .................... | .................... | .................... | |
| K | 6222 | hCoV-19/Gambia/NPHL-1645/2020\|EPI_ISL_561236\|2020- | .................... | .................... | .................... | .................... | .................... | .................... | |
| K | 6223 | hCoV-19/Gambia/NPHL-1711/2020\|EPI_ISL_561237\|2020- | .................... | .................... | .................... | .................... | .................... | .................... | |
| K | 6224 | hCoV-19/Gambia/NPHL-1746/2020\|EPI_ISL_561238\|2020- | .................... | .................... | .................... | .................... | .................... | .................... | |
| K | 6225 | hCoV-19/Gambia/NPHL-1921/2020\|EPI_ISL_561239\|2020 | .................... | .................... | .................... | .................... | .................... | .................... | |
| K | 6226 | hCoV-19/Gambia/NPHL-1968/2020\|EPI_ISL_561240\|2020- | .................... | .................... | .................... | .................... | .................... | .................... | |
| K | 6227 | hCoV-19/Gambia/NPHL-2037/2020\|EPI_ISL_561241\|2020- | .................... | .................... | .................... | .................... | .................... | .................... | |
| K | 6229 | hCoV-19/Gambia/NPHL-2344/2020\|EPI_ISL_561243\|2020 | .................... | .................... | .................... | .................... | .................... | .................... | |
| K | 6230 | hCoV-19/Gambia/NPHL-2374/2020\|EPI_ISL_561244\|2020 | .................... | .................... | .................... | .................... | .................... | .................... | |
| K | 6231 | hCoV-19/Gambia/NPHL-242/2020\|EPI_ISL_561245\|2020 | .................... | .................... | .................... | .................... | .................... | .................... | |
| K | 6232 | hCoV-19/Gambia/NPHL-244/2020\|EPI_ISL_561247\|2020 | .................... | .................... | .................... | .................... | .................... | .................... | |
| K | 6233 | hCoV-19/Gambia/NPHL-2443/2020\|EPI_ISL_561248\|2020 | .................... | .................... | .................... | .................... | .................... | .................... | |
| K | 6234 | hCoV-19/Gambia/NPHL-2444/2020\|EPI_ISL_561249\|2020- | .................... | .................... | .................... | .................... | .................... | .................... | |
| K | 6236 | hCoV-19/Australia/VIC13769/2020\|EPI_ISL_563830\|202 | .................... | .................... | .................... | .................... | .................... | .................... | |
| K | 6237 | hCoV-19/Australia/VIC13774/2020\|EPI_ISL_563831\|202 | .................... | .................... | .................... | .................... | NN.................. | .................... | |
| K | 6238 | hCoV-19/Australia/VIC13777/2020\|EPI_ISL_563832\|202 | .................... | .................... | .................... | .................... | .................... | .................... | |
| K | 6239 | hCoV-19/Australia/VIC13780/2020\|EPI_ISL_563833\|202 | .................... | .................... | .................... | .................... | .................... | .................... | |
| K | 6240 | hCoV-19/Australia/VIC13782/2020\|EPI_ISL_563834\|202 | .................... | .................... | .................... | .................... | .................... | .................... | |
| K | 6241 | hCoV-19/Australia/VIC13784/2020\|EPI_ISL_563835\|202 | .................... | .................... | .................... | .................... | .................... | .................... | |
| K | 6242 | hCoV-19/Australia/VIC13786/2020\|EPI_ISL_563836\|202 | .................... | .................... | .................... | .................... | .................... | .................... | |
| K | 6243 | hCoV-19/Australia/VIC13789/2020\|EPI_ISL_563839\|202 | .................... | .................... | .................... | .................... | .................... | .................... | |
| K | 6244 | hCoV-19/Australia/VIC13838/2020\|EPI_ISL_563851\|202 | .................... | .................... | .................... | .................... | .................... | .................... | |
| K | 6245 | hCoV-19/Australia/VIC13841/2020\|EPI_ISL_563852\|202 | .................... | .................... | .................... | .................... | .................... | .................... | |
| K | 6246 | hCoV-19/Australia/VIC13845/2020\|EPI_ISL_563853\|202 | .................... | .................... | .................... | .................... | .................... | .................... | |
| K | 6247 | hCoV-19/Australia/VIC13851/2020\|EPI_ISL_563854\|202 | .................... | .................... | .................... | .................... | .................... | .................... | |
| K | 6248 | hCoV-19/Australia/VIC13852/2020\|EPI_ISL_563855\|202 | .................... | .................... | .................... | .................... | .................... | .................... | |
| K | 6249 | hCoV-19/Australia/VIC13853/2020\|EPI_ISL_563856\|202 | .................... | .................... | .................... | .................... | .................... | .................... | |
| K | 6250 | hCoV-19/Australia/VIC13857/2020\|EPI_ISL_563857\|202 | .................... | .................... | .................... | .................... | .................... | .................... | |
| K | 6251 | hCoV-19/Australia/VIC13867/2020\|EPI_ISL_563858\|202 | .................... | .................... | .................... | .................... | .................... | .................... | |
| K | 6252 | hCoV-19/Australia/VIC13869/2020\|EPI_ISL_563859\|202 | .................... | .................... | .................... | .................... | .................... | .................... | |
| K | 6253 | hCoV-19/Australia/VIC13870/2020\|EPI_ISL_563860\|202 | .................... | .................... | .................... | .................... | .................... | .................... | |
| K | 6254 | hCoV-19/Australia/VIC13871/2020\|EPI_ISL_563861\|202 | .................... | .................... | .................... | .................... | .................... | .................... | |
| K | 6255 | hCoV-19/Australia/VIC13878/2020\|EPI_ISL_563862\|202 | .................... | .................... | .................... | .................... | .................... | .................... | |
| K | 6256 | hCoV-19/Australia/VIC13881/2020\|EPI_ISL_563863\|202 | .................... | .................... | .................... | .................... | .................... | .................... | |
| K | 6257 | hCoV-19/Australia/VIC13888/2020\|EPI_ISL_563864\|202 | .................... | .................... | .................... | .................... | .................... | .................... | |
| K | 6260 | hCoV-19/Australia/VIC13891/2020\|EPI_ISL_563867\|202 | .................... | .................... | .................... | .................... | .................... | .................... | |
| K | 6261 | hCoV-19/Australia/VIC13894/2020\|EPI_ISL_563868\|202 | .................... | .................... | .................... | .................... | .................... | .................... | |
| K | 6262 | hCoV-19/Australia/VIC13895/2020\|EPI_ISL_563869\|202 | .................... | .................... | .................... | .................... | .................... | .................... | |
| K | 6263 | hCoV-19/Australia/VIC13896/2020\|EPI_ISL_563870\|202 | .................... | .................... | .................... | .................... | .................... | .................... | |
| K | 6264 | hCoV-19/Australia/VIC13898/2020\|EPI_ISL_563871\|202 | .................... | .................... | .................... | .................... | .................... | .................... | |
| K | 6265 | hCoV-19/Australia/VIC13900/2020\|EPI_ISL_563872\|202 | .................... | .................... | .................... | .................... | ..........W......... | .................... | |
| K | 6266 | hCoV-19/Australia/VIC13901/2020\|EPI_ISL_563873\|202 | .................... | .................... | .................... | .................... | .................... | .................... | |
| K | 6267 | hCoV-19/Australia/VIC13902/2020\|EPI_ISL_563874\|202 | .................... | .................... | .................... | .................... | .................... | .................... | |
| K | 6268 | hCoV-19/Australia/VIC13903/2020\|EPI_ISL_563875\|202 | .................... | .................... | .................... | .................... | .................... | .................... | |
| K | 6269 | hCoV-19/Australia/VIC13904/2020\|EPI_ISL_563876\|202 | .................... | .................... | .................... | .................... | .................... | .................... | |
| K | 6270 | hCoV-19/Australia/VIC13905/2020\|EPI_ISL_563877\|202 | .................... | .................... | .................... | .................... | .................... | .................... | |
| K | 6271 | hCoV-19/Australia/VIC13908/2020\|EPI_ISL_563878\|202 | .................... | .................... | .................... | .................... | .................... | .................... | |
| K | 6273 | hCoV-19/Australia/VIC13911/2020\|EPI_ISL_563880\|202 | .................... | .................... | .................... | .................... | .................... | .................... | |
| K | 6274 | hCoV-19/Australia/VIC13918/2020\|EPI_ISL_563881\|202 | .................... | .................... | .................... | .................... | .................... | .................... | |
| K | 6275 | hCoV-19/Australia/VIC13919/2020\|EPI_ISL_563882\|202 | .................... | .................... | .................... | .................... | .................... | .................... | |
| K | 6276 | hCoV-19/Australia/VIC13923/2020\|EPI_ISL_563883\|202 | .................... | .................... | .................... | .................... | NNN................. | .................... | 1 |
| K | 6277 | hCoV-19/Australia/VIC13926/2020\|EPI_ISL_563884\|202 | .................... | .................... | .................... | .................... | ...K................ | .................... | |
| K | 6279 | hCoV-19/Australia/VIC13940/2020\|EPI_ISL_563886\|202 | .................... | .................... | .................... | .................... | N................... | .................... | |
| K | 6280 | hCoV-19/Australia/VIC13942/2020\|EPI_ISL_563887\|202 | .................... | .................... | .................... | .................... | N................... | .................... | |
| K | 6283 | hCoV-19/Australia/VIC13950/2020\|EPI_ISL_563890\|202 | .................... | .................... | .................... | .................... | .................... | .................... | |
| K | 6286 | hCoV-19/Australia/VIC13959/2020\|EPI_ISL_563893\|202 | .................... | .................... | .................... | .................... | .................... | .................... | |
| K | 6293 | hCoV-19/Australia/VIC6902/2020\|EPI_ISL_564126\|2020 | .................... | .................... | .................... | .................... | .................... | .................... | |
| K | 6294 | hCoV-19/Australia/VIC6903/2020\|EPI_ISL_564127\|2020 | .................... | .................... | .................... | .................... | .................... | .................... | |
| K | 6295 | hCoV-19/Australia/VIC6904/2020\|EPI_ISL_564128\|2020 | .................... | .................... | .................... | .................... | .................... | .................... | |
| K | 6296 | hCoV-19/Australia/VIC6905/2020\|EPI_ISL_564129\|2020 | .................... | .................... | .................... | .................... | .................... | .................... | |
| K | 6297 | hCoV-19/Australia/VIC6908/2020\|EPI_ISL_564130\|2020 | .................... | .................... | .................... | .................... | .................... | .................... | |
| K | 6298 | hCoV-19/Australia/VIC6909/2020\|EPI_ISL_564131\|2020 | .................... | .................... | .................... | .................... | .................... | .................... | |
| K | 6299 | hCoV-19/Australia/VIC6911/2020\|EPI_ISL_564132\|2020 | .................... | .................... | .................... | .................... | .................... | .................... | |
| K | 6300 | hCoV-19/Australia/VIC6912/2020\|EPI_ISL_564133\|2020 | .................... | .................... | .................... | .................... | .................... | .................... | |
| K | 6301 | hCoV-19/Australia/VIC6913/2020\|EPI_ISL_564134\|2020 | .................... | .................... | .................... | .................... | .................... | .................... | |
| K | 6302 | hCoV-19/Australia/VIC6914/2020\|EPI_ISL_564135\|2020 | .................... | .................... | .................... | .................... | .................... | .................... | |
| K | 6303 | hCoV-19/Australia/VIC6915/2020\|EPI_ISL_564136\|2020 | .................... | .................... | .................... | .................... | .................... | .................... | |
| K | 6304 | hCoV-19/Australia/VIC6917/2020\|EPI_ISL_564137\|2020 | .................... | .................... | .................... | .................... | .................... | .................... | |
| K | 6305 | hCoV-19/Australia/VIC6918/2020\|EPI_ISL_564138\|2020 | .................... | .................... | .................... | .................... | .................... | .................... | |
| K | 6306 | hCoV-19/Australia/VIC6919/2020\|EPI_ISL_564139\|2020 | .................... | .................... | .................... | .................... | .................... | .................... | |
| K | 6307 | hCoV-19/Australia/VIC6920/2020\|EPI_ISL_564140\|2020 | .................... | .................... | .................... | .................... | .................... | .................... | |
| K | 6308 | hCoV-19/Australia/VIC6921/2020\|EPI_ISL_564141\|2020 | .................... | .................... | .................... | .................... | .................... | .................... | |
| K | 6309 | hCoV-19/Australia/VIC6922/2020\|EPI_ISL_564142\|2020 | .................... | .................... | .................... | .................... | .................... | .................... | |
| K | 6310 | hCoV-19/Australia/VIC6923/2020\|EPI_ISL_564143\|2020 | .................... | .................... | .................... | .................... | .................... | .................... | |
| K | 6311 | hCoV-19/Australia/VIC6924/2020\|EPI_ISL_564144\|2020 | .................... | .................... | .................... | .................... | .................... | .................... | |
| K | 6312 | hCoV-19/Australia/VIC6925/2020\|EPI_ISL_564145\|2020 | .................... | .................... | .................... | .................... | .................... | .................... | |

| | | | | | | | |
|---|---|---|---|---|---|---|---|
| K | 6313 hCoV-19/Australia/VIC6927/2020\|EPI_ISL_564146\|2020 | ···················· | ···················· | ····················· | ···················· | ···················· | ···················· |
| K | 6314 hCoV-19/Australia/VIC6928/2020\|EPI_ISL_564147\|2020 | ···················· | ···················· | ····················· | ···················· | ···················· | ···················· |
| K | 6315 hCoV-19/Australia/VIC6929/2020\|EPI_ISL_564148\|2020 | ···················· | ···················· | ····················· | ···················· | ···················· | ···················· |
| K | 6316 hCoV-19/Australia/VIC6932/2020\|EPI_ISL_564149\|2020 | ···················· | ···················· | ····················· | ···················· | ···················· | ···················· |
| K | 6317 hCoV-19/Australia/VIC6934/2020\|EPI_ISL_564151\|2020 | ···················· | ···················· | ····················· | ···················· | ···················· | ···················· |
| K | 6318 hCoV-19/Australia/VIC6935/2020\|EPI_ISL_564152\|2020 | ···················· | ···················· | ····················· | ···················· | ···················· | ···················· |
| K | 6319 hCoV-19/Australia/VIC6936/2020\|EPI_ISL_564153\|2020 | ···················· | ···················· | ····················· | ···················· | ···················· | ···················· |
| K | 6320 hCoV-19/Australia/VIC6938/2020\|EPI_ISL_564155\|2020 | ···················· | ···················· | ····················· | ···················· | ···················· | ···················· |
| K | 6321 hCoV-19/Australia/VIC6939/2020\|EPI_ISL_564156\|2020 | ···················· | ···················· | ····················· | ···················· | ···················· | ···················· |
| K | 6322 hCoV-19/Australia/VIC6941/2020\|EPI_ISL_564157\|2020 | ···················· | ···················· | ····················· | ···················· | ···················· | ···················· |
| K | 6323 hCoV-19/Australia/VIC6942/2020\|EPI_ISL_564158\|2020 | ···················· | ···················· | ····················· | ···················· | ···················· | ···················· |
| K | 6324 hCoV-19/Australia/VIC6945/2020\|EPI_ISL_564159\|2020 | ···················· | ···················· | ····················· | ···················· | ···················· | ···················· |
| K | 6327 hCoV-19/Australia/VIC6952/2020\|EPI_ISL_564162\|2020 | ···················· | ···················· | ····················· | ···················· | ···················· | ···················· |
| K | 6328 hCoV-19/Australia/VIC6960/2020\|EPI_ISL_564163\|2020 | ···················· | ···················· | ····················· | ···················· | ···················· | ···················· |
| K | 6330 hCoV-19/Australia/VIC6966/2020\|EPI_ISL_564165\|2020 | ···················· | ···················· | ····················· | ···················· | ···················· | ···················· |
| K | 6331 hCoV-19/Australia/VIC6967/2020\|EPI_ISL_564166\|2020 | ···················· | ···················· | ····················· | ···················· | ···················· | ···················· |
| K | 6332 hCoV-19/Australia/VIC6968/2020\|EPI_ISL_564167\|2020 | ···················· | ···················· | ····················· | ···················· | ···················· | ···················· |
| K | 6333 hCoV-19/Australia/VIC6969/2020\|EPI_ISL_564168\|2020 | ···················· | ···················· | ····················· | ···················· | ···················· | ···················· |
| K | 6334 hCoV-19/Australia/VIC6972/2020\|EPI_ISL_564169\|2020 | ···················· | ···················· | ····················· | ···················· | ···················· | ···················· |
| K | 6335 hCoV-19/Australia/VIC9235/2020\|EPI_ISL_565457\|2020 | ···················· | ···················· | ····················· | ···················· | ···················· | ···················· |
| K | 6336 hCoV-19/Australia/VIC9238/2020\|EPI_ISL_565458\|2020 | ···················· | ···················· | ····················· | ···················· | ···················· | ···················· |
| K | 6337 hCoV-19/Australia/VIC9239/2020\|EPI_ISL_565459\|2020 | ···················· | ···················· | ····················· | ···················· | ···················· | ···················· |
| K | 6338 hCoV-19/Australia/VIC9240/2020\|EPI_ISL_565460\|2020 | ···················· | ···················· | ····················· | ···················· | ···················· | ···················· |
| K | 6339 hCoV-19/Australia/VIC9242/2020\|EPI_ISL_565461\|2020 | ···················· | ···················· | ····················· | ···················· | ···················· | ···················· |
| K | 6340 hCoV-19/Australia/VIC9243/2020\|EPI_ISL_565462\|2020 | ···················· | ···················· | ····················· | ···················· | ···················· | ···················· |
| K | 6341 hCoV-19/Australia/VIC9246/2020\|EPI_ISL_565463\|2020 | ···················· | ···················· | ····················· | ···················· | ···················· | ···················· |
| K | 6342 hCoV-19/Australia/VIC9248/2020\|EPI_ISL_565464\|2020 | ···················· | ···················· | ····················· | ···················· | ···················· | ···················· |
| K | 6343 hCoV-19/Australia/VIC9250/2020\|EPI_ISL_565465\|2020 | ···················· | ···················· | ····················· | ···················· | ···················· | ···················· |
| K | 6344 hCoV-19/Australia/VIC9251/2020\|EPI_ISL_565466\|2020 | ···················· | ···················· | ····················· | ···················· | ···················· | ···················· |
| K | 6345 hCoV-19/Australia/VIC9252/2020\|EPI_ISL_565467\|2020 | ···················· | ···················· | ····················· | ···················· | ···················· | ···················· |
| K | 6346 hCoV-19/Australia/VIC9253/2020\|EPI_ISL_565468\|2020 | ···················· | ···················· | ····················· | ···················· | ···················· | ···················· |
| K | 6347 hCoV-19/Australia/VIC9254/2020\|EPI_ISL_565469\|2020 | ···················· | ···················· | ····················· | ···················· | ···················· | ···················· |
| K | 6348 hCoV-19/Australia/VIC9255/2020\|EPI_ISL_565470\|2020 | ···················· | ···················· | ····················· | ···················· | ···················· | ···················· |
| K | 6349 hCoV-19/Australia/VIC9256/2020\|EPI_ISL_565471\|2020 | ···················· | ···················· | ····················· | ···················· | ···················· | ···················· |
| K | 6350 hCoV-19/Australia/VIC9257/2020\|EPI_ISL_565472\|2020 | ···················· | ···················· | ····················· | ···················· | ···················· | ···················· |
| K | 6351 hCoV-19/Australia/VIC9258/2020\|EPI_ISL_565473\|2020 | ···················· | ···················· | ····················· | ···················· | ···················· | ···················· |
| K | 6352 hCoV-19/Australia/VIC9260/2020\|EPI_ISL_565474\|2020 | ···················· | ···················· | ····················· | ···················· | ···················· | ···················· |
| K | 6353 hCoV-19/Australia/VIC9264/2020\|EPI_ISL_565475\|2020 | ···················· | ···················· | ····················· | ···················· | ···················· | ···················· |
| K | 6354 hCoV-19/Australia/VIC9266/2020\|EPI_ISL_565476\|2020 | ···················· | ···················· | ····················· | ···················· | ···················· | ···················· |
| K | 6355 hCoV-19/Australia/VIC9267/2020\|EPI_ISL_565477\|2020 | ···················· | ···················· | ····················· | ···················· | ···················· | ···················· |
| K | 6356 hCoV-19/Australia/VIC9274/2020\|EPI_ISL_565478\|2020 | ···················· | ···················· | ····················· | ···················· | ···················· | ···················· |
| K | 6357 hCoV-19/Australia/VIC9277/2020\|EPI_ISL_565479\|2020 | ···················· | ···················· | ····················· | ···················· | ···················· | ···················· |
| K | 6358 hCoV-19/Australia/VIC9283/2020\|EPI_ISL_565482\|2020 | ···················· | ···················· | ····················· | ···················· | ···················· | ···················· |
| K | 6359 hCoV-19/Australia/VIC9284/2020\|EPI_ISL_565483\|2020 | ···················· | ···················· | ····················· | ···················· | ···················· | ···················· |
| K | 6360 hCoV-19/Australia/VIC9287/2020\|EPI_ISL_565485\|2020 | ···················· | ···················· | ····················· | ···················· | ···················· | ···················· |
| K | 6361 hCoV-19/Australia/VIC9288/2020\|EPI_ISL_565486\|2020 | ···················· | ···················· | ····················· | ···················· | ···················· | ···················· |
| K | 6362 hCoV-19/Australia/VIC9290/2020\|EPI_ISL_565487\|2020 | ···················· | ···················· | ····················· | ···················· | ···················· | ···················· |
| K | 6363 hCoV-19/Australia/VIC9292/2020\|EPI_ISL_565488\|2020 | ···················· | ···················· | ····················· | ···················· | ···················· | ···················· |
| K | 6364 hCoV-19/Australia/VIC9293/2020\|EPI_ISL_565489\|2020 | ···················· | ···················· | ····················· | ···················· | ···················· | ···················· |
| K | 6365 hCoV-19/Australia/VIC9294/2020\|EPI_ISL_565490\|2020 | ···················· | ···················· | ····················· | ···················· | ···················· | ···················· |
| K | 6366 hCoV-19/Australia/VIC9295/2020\|EPI_ISL_565491\|2020 | ···················· | ···················· | ····················· | ···················· | ···················· | ···················· |
| K | 6367 hCoV-19/Australia/VIC9296/2020\|EPI_ISL_565492\|2020 | ···················· | ···················· | ····················· | ···················· | ···················· | ···················· |
| K | 6368 hCoV-19/Australia/VIC9298/2020\|EPI_ISL_565493\|2020 | ···················· | ···················· | ····················· | ···················· | ···················· | ···················· |
| K | 6369 hCoV-19/Australia/VIC9299/2020\|EPI_ISL_565494\|2020 | ···················· | ···················· | ····················· | ···················· | ···················· | ···················· |
| K | 6370 hCoV-19/Australia/VIC9300/2020\|EPI_ISL_565495\|2020 | ···················· | ···················· | ····················· | ···················· | ···················· | ···················· |
| K | 6371 hCoV-19/Australia/VIC9302/2020\|EPI_ISL_565496\|2020 | ···················· | ···················· | ····················· | ···················· | ···················· | ···················· |
| K | 6372 hCoV-19/Australia/VIC9303/2020\|EPI_ISL_565497\|2020 | ···················· | ···················· | ····················· | ···················· | ···················· | ···················· |
| K | 6373 hCoV-19/Australia/VIC9306/2020\|EPI_ISL_565498\|2020 | ···················· | ···················· | ····················· | ···················· | ···················· | ···················· |
| K | 6374 hCoV-19/Australia/VIC9311/2020\|EPI_ISL_565499\|2020 | ···················· | ···················· | ····················· | ···················· | ···················· | ···················· |
| K | 6375 hCoV-19/England/MILK-9C3598/2020\|EPI_ISL_566789\|20 | ···················· | ···················· | ····················· | ···················· | ···················· | ···················· |
| K | 6376 hCoV-19/England/MILK-9C347D/2020\|EPI_ISL_566790\|20 | ···················· | ·A·················· | ····················· | ···················· | ···················· | ···A················ |
| K | 6377 hCoV-19/England/MILK-9C3370/2020\|EPI_ISL_566792\|20 | ···················· | ···················· | ····················· | ···················· | ···················· | ···················· |
| K | 6378 hCoV-19/England/MILK-9C3F21/2020\|EPI_ISL_566793\|20 | ···················· | ···················· | ····················· | ···················· | ······T············· | ···················· |
| K | 6379 hCoV-19/England/MILK-9C3D45/2020\|EPI_ISL_566794\|20 | ···················· | ···················· | ····················· | ···················· | ···················· | ···················· |
| K | 6380 hCoV-19/England/MILK-9C4658/2020\|EPI_ISL_566796\|20 | ···················· | ···················· | ····················· | ···················· | ···················· | ···················· |
| K | 6381 hCoV-19/England/MILK-9C3C1B/2020\|EPI_ISL_566797\|20 | ···················· | ···················· | ····················· | ···················· | ···················· | ···················· |
| K | 6382 hCoV-19/England/MILK-9C46EF/2020\|EPI_ISL_566798\|20 | ···················· | ···················· | ····················· | ···················· | ···················· | ···················· |
| K | 6383 hCoV-19/France/PAC-IHU-0945/2020\|EPI_ISL_569000\|20 | ···················· | ····A··············· | ····················· | ···················· | ···················· | ···················· |
| K | 6384 hCoV-19/France/PAC-IHU-0948/2020\|EPI_ISL_569002\|20 | ···················· | ····A··············· | ····················· | ···················· | ···················· | ···················· |
| K | 6385 hCoV-19/France/PAC-IHU-0949/2020\|EPI_ISL_569003\|20 | ···················· | ····A··············· | ····················· | ···················· | ···················· | ···················· |
| K | 6386 hCoV-19/France/PAC-IHU-0950/2020\|EPI_ISL_569004\|20 | ···················· | ····A··············· | ····················· | ···················· | ···················· | ···················· |
| K | 6387 hCoV-19/France/PAC-IHU-0953/2020\|EPI_ISL_569005\|20 | ···················· | ···················· | ····················· | ···················· | ···················· | ···················· |
| K | 6388 hCoV-19/France/PAC-IHU-0954/2020\|EPI_ISL_569006\|20 | ···················· | ···················· | ····················· | ···················· | ···················· | ···················· |
| K | 6389 hCoV-19/France/PAC-IHU-0956/2020\|EPI_ISL_569008\|20 | ···················· | ····A··············· | ····················· | ···················· | ···················· | ···················· |
| K | 6390 hCoV-19/France/PAC-IHU-0957/2020\|EPI_ISL_569009\|20 | ···················· | ····A··············· | ····················· | ···················· | ···················· | ···················· |
| K | 6391 hCoV-19/France/PAC-IHU-0958/2020\|EPI_ISL_569010\|20 | ···················· | ····A··············· | ····················· | ···················· | ···················· | ···················· |
| K | 6392 hCoV-19/France/PAC-IHU-0959/2020\|EPI_ISL_569011\|20 | ···················· | ····A··············· | ····················· | ···················· | ···················· | ···················· |
| K | 6393 hCoV-19/France/PAC-IHU-0962/2020\|EPI_ISL_569012\|20 | ···················· | ···················· | ····················· | ···················· | ···················· | ···················· |
| K | 6394 hCoV-19/France/PAC-IHU-0963/2020\|EPI_ISL_569013\|20 | ···················· | ····A··············· | ····················· | ···················· | ···················· | ···················· |
| K | 6395 hCoV-19/France/PAC-IHU-0964/2020\|EPI_ISL_569014\|20 | ···················· | ····A··············· | ····················· | ···················· | ···················· | ···················· |

| | | | | | | | | |
|---|---|---|---|---|---|---|---|---|
| K | 6396 | hCoV-19/France/PAC-IHU-0965/2020\|EPI_ISL_569015\|20 | ···················· | ···················· | ···················· | ···················· | ···················· | ···················· |
| K | 6397 | hCoV-19/France/PAC-IHU-0966/2020\|EPI_ISL_569016\|20 | ···················· | ····A··············· | ···················· | ···················· | ···················· | ···················· |
| K | 6398 | hCoV-19/France/PAC-IHU-0967/2020\|EPI_ISL_569017\|20 | ···················· | ·A·················· | ···················· | ···················· | ···················· | ···A················ |
| K | 6399 | hCoV-19/France/PAC-IHU-0970/2020\|EPI_ISL_569018\|20 | ···················· | ····A··············· | ···················· | ···················· | ···················· | ···················· |
| K | 6400 | hCoV-19/France/PAC-IHU-0971/2020\|EPI_ISL_569019\|20 | ···················· | ···················· | ···················· | ···················· | ···················· | ···················· |
| K | 6401 | hCoV-19/England/GSTT-5631/2020\|EPI_ISL_560695\|2020 | ···················· | ···················· | ···················· | ···················· | ···················· | ···················· |
| K | 6402 | hCoV-19/England/GSTT-5633/2020\|EPI_ISL_560696\|2020 | ···················· | ···················· | ···················· | ···················· | ···················· | ···················· |
| K | 6403 | hCoV-19/England/GSTT-5646/2020\|EPI_ISL_560697\|2020 | ···················· | ···················· | ···················· | ···················· | ···················· | ···················· |
| K | 6404 | hCoV-19/England/GSTT-5651/2020\|EPI_ISL_560698\|2020 | ···················· | ···················· | ···················· | ···················· | ···················· | ···················· |
| K | 6405 | hCoV-19/England/GSTT-5655/2020\|EPI_ISL_560699\|2020 | ···················· | ···················· | ···················· | ···················· | ···················· | ···················· |
| K | 6406 | hCoV-19/Australia/VIC6933/2020\|EPI_ISL_564150\|2020 | ···················· | ···················· | ···················· | ···················· | ···················· | ···················· |
| K | 6407 | hCoV-19/Australia/VIC6937/2020\|EPI_ISL_564154\|2020 | ···················· | ···················· | ···················· | ···················· | ···················· | ···················· |
| K | 6408 | hCoV-19/Australia/VIC6973/2020\|EPI_ISL_564170\|2020 | ···················· | ···················· | ···················· | ···················· | ···················· | ···················· |
| K | 6409 | hCoV-19/Australia/VIC6974/2020\|EPI_ISL_564171\|2020 | ···················· | ···················· | ···················· | ···················· | ···················· | ···················· |
| K | 6410 | hCoV-19/Australia/VIC6975/2020\|EPI_ISL_564172\|2020 | ···················· | ···················· | ···················· | ···················· | ···················· | ···················· |
| K | 6411 | hCoV-19/Australia/VIC6976/2020\|EPI_ISL_564173\|2020 | ···················· | ···················· | ···················· | ···················· | ···················· | ···················· |
| K | 6412 | hCoV-19/Australia/VIC6977/2020\|EPI_ISL_564174\|2020 | ···················· | ···················· | ···················· | ···················· | ···················· | ···················· |
| K | 6413 | hCoV-19/Australia/VIC6978/2020\|EPI_ISL_564175\|2020 | ···················· | ···················· | ···················· | ···················· | ···················· | ···················· |
| K | 6414 | hCoV-19/Australia/VIC6979/2020\|EPI_ISL_564176\|2020 | ···················· | ···················· | ···················· | ···················· | ···················· | ···················· |
| K | 6415 | hCoV-19/Australia/VIC6980/2020\|EPI_ISL_564177\|2020 | ···················· | ···················· | ···················· | ···················· | ···················· | ···················· |
| K | 6420 | hCoV-19/Australia/VIC6987/2020\|EPI_ISL_564182\|2020 | ···················· | ···················· | ···················· | ···················· | ···················· | ···················· |
| K | 6421 | hCoV-19/Australia/VIC6989/2020\|EPI_ISL_564183\|2020 | ···················· | ···················· | ···K················ | ···················· | ···················· | ···················· |
| K | 6422 | hCoV-19/Australia/VIC6990/2020\|EPI_ISL_564184\|2020 | ···················· | ···················· | ···················· | ···················· | ···················· | ···················· |
| K | 6426 | hCoV-19/Australia/VIC6994/2020\|EPI_ISL_564188\|2020 | ···················· | ···················· | ···················· | ···················· | ···················· | ···················· |
| K | 6427 | hCoV-19/Australia/VIC6996/2020\|EPI_ISL_564189\|2020 | ···················· | ···················· | ···················· | ···················· | ···················· | ···················· |
| K | 6428 | hCoV-19/Australia/VIC6998/2020\|EPI_ISL_564190\|2020 | ···················· | ···················· | ···················· | ···················· | ···················· | ···················· |
| K | 6429 | hCoV-19/Australia/VIC6999/2020\|EPI_ISL_564191\|2020 | ···················· | ···················· | ···················· | ···················· | ···················· | ···················· |
| K | 6432 | hCoV-19/Australia/VIC7005/2020\|EPI_ISL_564194\|2020 | ···················· | ···················· | ···················· | ···················· | ···················· | ···················· |
| K | 6433 | hCoV-19/Australia/VIC7006/2020\|EPI_ISL_564195\|2020 | ···················· | ···················· | ···················· | ···················· | ···················· | ···················· |
| K | 6435 | hCoV-19/Australia/VIC7012/2020\|EPI_ISL_564197\|2020 | ···················· | ···················· | ···················· | ···················· | ···················· | ···················· |
| K | 6438 | hCoV-19/Australia/VIC7027/2020\|EPI_ISL_564200\|2020 | ···················· | ···················· | ···················· | ···················· | ···················· | ···················· |
| K | 6439 | hCoV-19/Australia/VIC7029/2020\|EPI_ISL_564201\|2020 | ···················· | ······A············· | ···K················ | ···················· | ···················· | ···················· |
| K | 6440 | hCoV-19/Australia/VIC7034/2020\|EPI_ISL_564202\|2020 | ···················· | ···················· | ···················· | ···················· | ···················· | ···················· |
| K | 6443 | hCoV-19/Australia/VIC7041/2020\|EPI_ISL_564205\|2020 | ···················· | ···················· | ···················· | ···················· | ···················· | ···················· |
| K | 6444 | hCoV-19/Australia/VIC7044/2020\|EPI_ISL_564206\|2020 | ···················· | ···················· | ···················· | ···················· | ···················· | ···················· |
| K | 6445 | hCoV-19/Australia/VIC7046/2020\|EPI_ISL_564207\|2020 | ···················· | ···················· | ···················· | ···················· | ···················· | ···················· |
| K | 6447 | hCoV-19/Australia/VIC7050/2020\|EPI_ISL_564209\|2020 | ···················· | ···················· | ···K················ | ···················· | ···················· | ···················· |
| K | 6448 | hCoV-19/Australia/VIC7053/2020\|EPI_ISL_564210\|2020 | ···················· | ···················· | ···················· | ···················· | ···················· | ···················· |
| K | 6450 | hCoV-19/Australia/VIC7055/2020\|EPI_ISL_564212\|2020 | ···················· | ···················· | ···················· | ···················· | ···················· | ···················· |
| K | 6451 | hCoV-19/Australia/VIC7066/2020\|EPI_ISL_564213\|2020 | ···················· | ···················· | ···················· | ···················· | ···················· | ···················· |
| K | 6452 | hCoV-19/Australia/VIC7067/2020\|EPI_ISL_564214\|2020 | ···················· | ···················· | ···················· | ···················· | ···················· | ···················· |
| K | 6455 | hCoV-19/Australia/VIC7074/2020\|EPI_ISL_564217\|2020 | ···················· | ···················· | ···················· | ···················· | ···················· | ···················· |
| K | 6456 | hCoV-19/Australia/VIC7079/2020\|EPI_ISL_564218\|2020 | ···················· | ···················· | ···················· | ···················· | ···················· | ···················· |
| K | 6457 | hCoV-19/Australia/VIC7080/2020\|EPI_ISL_564219\|2020 | ···················· | ···················· | ···················· | ···················· | ···················· | ···················· |
| K | 6458 | hCoV-19/Australia/VIC7081/2020\|EPI_ISL_564220\|2020 | ···················· | ···················· | ···················· | ···················· | ···················· | ·K·················· |
| K | 6459 | hCoV-19/Australia/VIC7082/2020\|EPI_ISL_564221\|2020 | ···················· | ···················· | ···················· | ···················· | ···················· | ···················· |
| K | 6460 | hCoV-19/Australia/VIC7084/2020\|EPI_ISL_564222\|2020 | ···················· | ···················· | ···················· | ···················· | ···················· | ···················· |
| K | 6461 | hCoV-19/Australia/VIC7089/2020\|EPI_ISL_564223\|2020 | ···················· | ···················· | ···················· | ···················· | ···················· | ···················· |
| K | 6462 | hCoV-19/Australia/VIC7090/2020\|EPI_ISL_564224\|2020 | ···················· | ···················· | ···················· | ···················· | ···················· | ···················· |
| K | 6463 | hCoV-19/Australia/VIC7091/2020\|EPI_ISL_564225\|2020 | ···················· | ···················· | ···················· | ···················· | ···················· | ···················· |
| K | 6464 | hCoV-19/Australia/VIC7092/2020\|EPI_ISL_564226\|2020 | ···················· | ···················· | ···················· | ···················· | ···················· | ···················· |
| K | 6465 | hCoV-19/Australia/VIC7095/2020\|EPI_ISL_564227\|2020 | ···················· | ···················· | ···················· | ···················· | ···················· | ···················· |
| K | 6466 | hCoV-19/Australia/VIC7097/2020\|EPI_ISL_564228\|2020 | ···················· | ···················· | ···················· | ···················· | ···················· | ···················· |
| K | 6467 | hCoV-19/Australia/VIC7098/2020\|EPI_ISL_564229\|2020 | ···················· | ···················· | ···················· | ···················· | ···················· | ···················· |
| K | 6468 | hCoV-19/Australia/VIC7099/2020\|EPI_ISL_564230\|2020 | ···················· | ···················· | ···················· | ···················· | ···················· | ···················· |
| K | 6469 | hCoV-19/Australia/VIC7100/2020\|EPI_ISL_564231\|2020 | ···················· | ···················· | ···················· | ···················· | ···················· | ···················· |
| K | 6470 | hCoV-19/Australia/VIC7101/2020\|EPI_ISL_564232\|2020 | ···················· | ···················· | ···················· | ···················· | ···················· | ···················· |
| K | 6471 | hCoV-19/Australia/VIC7103/2020\|EPI_ISL_564233\|2020 | ···················· | ···················· | ···················· | ···················· | ···················· | ···················· |
| K | 6472 | hCoV-19/Australia/VIC7105/2020\|EPI_ISL_564234\|2020 | ···················· | ···················· | ···················· | ···················· | ···················· | ···················· |
| K | 6473 | hCoV-19/Australia/VIC7106/2020\|EPI_ISL_564235\|2020 | ···················· | ···················· | ···················· | ···················· | ···················· | ···················· |
| K | 6474 | hCoV-19/Australia/VIC7108/2020\|EPI_ISL_564236\|2020 | ···················· | ···················· | ···················· | ···················· | ···················· | ···················· |
| K | 6475 | hCoV-19/Australia/VIC7111/2020\|EPI_ISL_564237\|2020 | ···················· | ···················· | ···················· | ···················· | ···················· | ···················· |
| K | 6476 | hCoV-19/Australia/VIC7113/2020\|EPI_ISL_564238\|2020 | ···················· | ···················· | ···················· | ···················· | ···················· | ···················· |
| K | 6477 | hCoV-19/Australia/VIC7114/2020\|EPI_ISL_564239\|2020 | ···················· | ···················· | ···················· | ···················· | ···················· | ···················· |
| K | 6478 | hCoV-19/Australia/VIC7115/2020\|EPI_ISL_564240\|2020 | ···················· | ···················· | ···················· | ···················· | ···················· | ···················· |
| K | 6479 | hCoV-19/Australia/VIC7116/2020\|EPI_ISL_564241\|2020 | ···················· | ····A··············· | ···················· | ···················· | ···················· | ···················· |
| K | 6480 | hCoV-19/Australia/VIC7117/2020\|EPI_ISL_564242\|2020 | ···················· | ···················· | ···················· | ···················· | ···················· | ···················· |
| K | 6481 | hCoV-19/Australia/VIC7118/2020\|EPI_ISL_564243\|2020 | ···················· | ···················· | ···················· | ···················· | ···················· | ···················· |
| K | 6482 | hCoV-19/Australia/VIC7124/2020\|EPI_ISL_564244\|2020 | ···················· | ···················· | ···················· | ···················· | ···················· | ···················· |
| K | 6483 | hCoV-19/Australia/VIC7125/2020\|EPI_ISL_564245\|2020 | ···················· | ···················· | ···················· | ···················· | ···················· | ···················· |
| K | 6484 | hCoV-19/Australia/VIC7126/2020\|EPI_ISL_564246\|2020 | ···················· | ···················· | ···················· | ···················· | ···················· | ···················· |
| K | 6485 | hCoV-19/Australia/VIC9278/2020\|EPI_ISL_565480\|2020 | ···················· | ···················· | ···················· | ···················· | ···················· | ···················· |
| K | 6486 | hCoV-19/Australia/VIC9279/2020\|EPI_ISL_565481\|2020 | ···················· | ···················· | ···················· | ···················· | ···················· | ···················· |
| K | 6487 | hCoV-19/Australia/VIC9285/2020\|EPI_ISL_565484\|2020 | ···················· | ···················· | ···················· | ···················· | ···················· | ···················· |
| K | 6488 | hCoV-19/Australia/VIC9312/2020\|EPI_ISL_565500\|2020 | ···················· | ···················· | ···················· | ···················· | ···················· | ···················· |
| K | 6489 | hCoV-19/Australia/VIC9313/2020\|EPI_ISL_565501\|2020 | ···················· | ···················· | ···················· | ···················· | ···················· | ···················· |
| K | 6490 | hCoV-19/Australia/VIC9314/2020\|EPI_ISL_565502\|2020 | ···················· | ···················· | ···················· | ···················· | ···················· | ···················· |
| K | 6491 | hCoV-19/Australia/VIC9318/2020\|EPI_ISL_565503\|2020 | ···················· | ···················· | ···················· | ···················· | ···················· | ···················· |
| K | 6492 | hCoV-19/Australia/VIC9319/2020\|EPI_ISL_565504\|2020 | ···················· | ···················· | ···················· | ···················· | ···················· | ···················· |
| K | 6493 | hCoV-19/Australia/VIC9321/2020\|EPI_ISL_565505\|2020 | ···················· | ···················· | ···················· | ···················· | ···················· | ···················· |

| | | | | | | | |
|---|---|---|---|---|---|---|---|
| K | 6494 hCoV-19/Australia/VIC9322/2020\|EPI_ISL_565506\|2020 | ···················· | ···················· | ···················· | ···················· | ···················· | ···················· |
| K | 6495 hCoV-19/Australia/VIC9323/2020\|EPI_ISL_565507\|2020 | ···················· | ···················· | ···················· | ···················· | ···················· | ···················· |
| K | 6496 hCoV-19/Australia/VIC9324/2020\|EPI_ISL_565508\|2020 | ···················· | ···················· | ···················· | ···················· | ···················· | ···················· |
| K | 6497 hCoV-19/Australia/VIC9333/2020\|EPI_ISL_565509\|2020 | ···················· | ···················· | ··K················· | ···················· | ···················· | ····N··············· |
| K | 6498 hCoV-19/Australia/VIC9343/2020\|EPI_ISL_565510\|2020 | ···················· | ···················· | ···················· | ···················· | ···················· | ···················· |
| K | 6499 hCoV-19/Australia/VIC9344/2020\|EPI_ISL_565511\|2020 | ···················· | ···················· | ···················· | ···················· | ···················· | ···················· |
| K | 6500 hCoV-19/Australia/VIC9345/2020\|EPI_ISL_565512\|2020 | ···················· | ·····A·············· | ···················· | ···················· | ···················· | ···················· |
| K | 6501 hCoV-19/Australia/VIC9346/2020\|EPI_ISL_565513\|2020 | ···················· | ···················· | ···················· | ···················· | ···················· | ···················· |
| K | 6502 hCoV-19/Australia/VIC9349/2020\|EPI_ISL_565514\|2020 | ···················· | ···················· | ···················· | ···················· | ···················· | ···················· |
| K | 6503 hCoV-19/Australia/VIC9350/2020\|EPI_ISL_565515\|2020 | ···················· | ···················· | ···················· | ···················· | ···················· | ···················· |
| K | 6504 hCoV-19/Australia/VIC9351/2020\|EPI_ISL_565516\|2020 | ···················· | ···················· | ···················· | ···················· | ···················· | ···················· |
| K | 6505 hCoV-19/Australia/VIC9353/2020\|EPI_ISL_565517\|2020 | ···················· | ···················· | ···················· | ···················· | ···················· | ···················· |
| K | 6506 hCoV-19/Australia/VIC9354/2020\|EPI_ISL_565518\|2020 | ···················· | ···················· | ···················· | ···················· | ···················· | ···················· |
| K | 6507 hCoV-19/Australia/VIC9356/2020\|EPI_ISL_565519\|2020 | ···················· | ···················· | ···················· | ···················· | ···················· | ···················· |
| K | 6508 hCoV-19/Australia/VIC9359/2020\|EPI_ISL_565520\|2020 | ···················· | ···················· | ···················· | ···················· | ···················· | ···················· |
| K | 6509 hCoV-19/Australia/VIC9363/2020\|EPI_ISL_565521\|2020 | ···················· | ···················· | ···················· | ···················· | ···················· | ···················· |
| K | 6510 hCoV-19/Australia/VIC9365/2020\|EPI_ISL_565522\|2020 | ···················· | ···················· | ···················· | ···················· | ···················· | ···················· |
| K | 6511 hCoV-19/Australia/VIC9366/2020\|EPI_ISL_565523\|2020 | ···················· | ···················· | ···················· | ···················· | ···················· | ···················· |
| K | 6512 hCoV-19/Australia/VIC9367/2020\|EPI_ISL_565524\|2020 | ···················· | ···················· | ···················· | ···················· | ···················· | ···················· |
| K | 6513 hCoV-19/Australia/VIC9368/2020\|EPI_ISL_565525\|2020 | ···················· | ···················· | ···················· | ···················· | ···················· | ···················· |
| K | 6514 hCoV-19/Australia/VIC9371/2020\|EPI_ISL_565526\|2020 | ···················· | ···················· | ···················· | ···················· | ···················· | ···················· |
| K | 6515 hCoV-19/Australia/VIC9374/2020\|EPI_ISL_565527\|2020 | ···················· | ···················· | ···················· | ···················· | ···················· | ···················· |
| K | 6516 hCoV-19/Australia/VIC9375/2020\|EPI_ISL_565528\|2020 | ···················· | ···················· | ···················· | ···················· | ···················· | ···················· |
| K | 6517 hCoV-19/Australia/VIC9376/2020\|EPI_ISL_565529\|2020 | ···················· | ···················· | ···················· | ···················· | ···················· | ···················· |
| K | 6518 hCoV-19/Australia/VIC9377/2020\|EPI_ISL_565530\|2020 | ···················· | ···················· | ···················· | ···················· | ···················· | ···················· |
| K | 6519 hCoV-19/Australia/VIC9378/2020\|EPI_ISL_565531\|2020 | ···················· | ···················· | ···················· | ···················· | ···················· | ···················· |
| K | 6520 hCoV-19/Australia/VIC9379/2020\|EPI_ISL_565532\|2020 | ···················· | ···················· | ···················· | ···················· | ···················· | ···················· |
| K | 6521 hCoV-19/Australia/VIC9382/2020\|EPI_ISL_565533\|2020 | ···················· | ···················· | ···················· | ···················· | ···················· | ···················· |
| K | 6522 hCoV-19/Australia/VIC9383/2020\|EPI_ISL_565534\|2020 | ···················· | ···················· | ···················· | ···················· | ···················· | ···················· |
| K | 6523 hCoV-19/Australia/VIC9389/2020\|EPI_ISL_565535\|2020 | ···················· | ···················· | ···················· | ···················· | ···················· | ···················· |
| K | 6524 hCoV-19/Australia/VIC9397/2020\|EPI_ISL_565536\|2020 | ···················· | ···················· | ···················· | ···················· | ···················· | ···················· |
| K | 6525 hCoV-19/Australia/VIC9403/2020\|EPI_ISL_565537\|2020 | ···················· | ···················· | ···················· | ···················· | ···················· | ···················· |
| K | 6526 hCoV-19/Australia/VIC9408/2020\|EPI_ISL_565538\|2020 | ···················· | ···················· | ···················· | ···················· | ···················· | ···················· |
| K | 6527 hCoV-19/Australia/VIC9409/2020\|EPI_ISL_565539\|2020 | ···················· | ···················· | ···················· | ···················· | ···················· | ···················· |
| K | 6528 hCoV-19/Australia/VIC9413/2020\|EPI_ISL_565540\|2020 | ···················· | ···················· | ···················· | ···················· | ···················· | ···················· |
| K | 6529 hCoV-19/Australia/VIC9416/2020\|EPI_ISL_565541\|2020 | ···················· | ···················· | ···················· | ···················· | ···················· | ···················· |
| K | 6530 hCoV-19/Australia/VIC9420/2020\|EPI_ISL_565542\|2020 | ···················· | ···················· | ···················· | ···················· | ···················· | ···················· |
| K | 6531 hCoV-19/Australia/VIC9424/2020\|EPI_ISL_565543\|2020 | ···················· | ···················· | ···················· | ···················· | ···················· | ···················· |
| K | 6532 hCoV-19/Australia/VIC9428/2020\|EPI_ISL_565544\|2020 | ···················· | ···················· | ···················· | ···················· | ···················· | ···················· |
| K | 6533 hCoV-19/Australia/VIC9430/2020\|EPI_ISL_565545\|2020 | ···················· | ···················· | ···················· | ···················· | ···················· | ···················· |
| K | 6534 hCoV-19/Australia/VIC9431/2020\|EPI_ISL_565546\|2020 | ···················· | ···················· | ···················· | ···················· | ···················· | ···················· |
| K | 6535 hCoV-19/Australia/VIC9433/2020\|EPI_ISL_565547\|2020 | ···················· | ···················· | ···················· | ···················· | ···················· | ···················· |
| K | 6536 hCoV-19/Australia/VIC9435/2020\|EPI_ISL_565548\|2020 | ···················· | ···················· | ···················· | ···················· | ···················· | ···················· |
| K | 6537 hCoV-19/Australia/VIC9436/2020\|EPI_ISL_565549\|2020 | ···················· | ···················· | ···················· | ···················· | ···················· | ···················· |
| K | 6538 hCoV-19/Australia/VIC9437/2020\|EPI_ISL_565550\|2020 | ···················· | ···················· | ···················· | ···················· | ···················· | ···················· |
| K | 6539 hCoV-19/Australia/VIC9438/2020\|EPI_ISL_565551\|2020 | ···················· | ···················· | ···················· | ···················· | ···················· | ···················· |
| K | 6540 hCoV-19/Australia/VIC9439/2020\|EPI_ISL_565552\|2020 | ···················· | ···················· | ···················· | ···················· | ···················· | ···················· |
| K | 6541 hCoV-19/Australia/VIC9440/2020\|EPI_ISL_565553\|2020 | ···················· | ···················· | ···················· | ···················· | ···················· | ···················· |
| K | 6542 hCoV-19/Australia/VIC9441/2020\|EPI_ISL_565554\|2020 | ···················· | ···················· | ···················· | ···················· | ···················· | ···················· |
| K | 6543 hCoV-19/Australia/VIC9443/2020\|EPI_ISL_565555\|2020 | ···················· | ···················· | ···················· | ···················· | ···················· | ···················· |
| K | 6544 hCoV-19/Australia/VIC9444/2020\|EPI_ISL_565556\|2020 | ···················· | ···················· | ···················· | ···················· | ···················· | ···················· |
| K | 6545 hCoV-19/Australia/VIC9452/2020\|EPI_ISL_565557\|2020 | ···················· | ···················· | ···················· | ···················· | ···················· | ···················· |
| K | 6547 hCoV-19/Australia/VIC9455/2020\|EPI_ISL_565559\|2020 | ···················· | ···················· | ···················· | ···················· | ···················· | ···················· |
| K | 6548 hCoV-19/Australia/VIC9456/2020\|EPI_ISL_565560\|2020 | ···················· | ···················· | ···················· | ···················· | ···················· | ···················· |
| K | 6549 hCoV-19/Australia/VIC9459/2020\|EPI_ISL_565561\|2020 | ···················· | ···················· | ···················· | ···················· | ···················· | ···················· |
| K | 6550 hCoV-19/Australia/VIC9460/2020\|EPI_ISL_565562\|2020 | ···················· | ···················· | ···················· | ···················· | ···················· | ···················· |
| K | 6553 hCoV-19/Australia/VIC9475/2020\|EPI_ISL_565565\|2020 | ···················· | ···················· | ···················· | ···················· | ···················· | ···················· |
| K | 6554 hCoV-19/Australia/VIC9476/2020\|EPI_ISL_565566\|2020 | ···················· | ···················· | ···················· | ···················· | ···················· | ···················· |
| K | 6555 hCoV-19/Australia/VIC9478/2020\|EPI_ISL_565567\|2020 | ···················· | ···················· | ···················· | ···················· | ···················· | ···················· |
| K | 6556 hCoV-19/Australia/VIC9486/2020\|EPI_ISL_565568\|2020 | ···················· | ···················· | ···················· | ···················· | ···················· | ···················· |
| K | 6557 hCoV-19/Australia/VIC9488/2020\|EPI_ISL_565569\|2020 | ···················· | ···················· | ···················· | ···················· | ···················· | ···················· |
| K | 6558 hCoV-19/Australia/VIC9489/2020\|EPI_ISL_565570\|2020 | ···················· | ···················· | ···················· | ···················· | ···············NN··· | ···················· |
| K | 6559 hCoV-19/Australia/VIC9490/2020\|EPI_ISL_565571\|2020 | ···················· | ···················· | ···················· | ···················· | ···················· | ···················· |
| K | 6560 hCoV-19/Australia/VIC9491/2020\|EPI_ISL_565572\|2020 | ···················· | ···················· | ···················· | ···················· | ···················· | ···················· |
| K | 6562 hCoV-19/Australia/VIC9494/2020\|EPI_ISL_565574\|2020 | ···················· | ···················· | ···················· | ···················· | ···················· | ···················· |
| K | 6563 hCoV-19/Australia/VIC9496/2020\|EPI_ISL_565575\|2020 | ···················· | ···················· | ···················· | ···················· | ···················· | ···················· |
| K | 6564 hCoV-19/Australia/VIC9499/2020\|EPI_ISL_565576\|2020 | ···················· | ···················· | ···················· | ···················· | ···················· | ···················· |
| K | 6565 hCoV-19/Australia/VIC9504/2020\|EPI_ISL_565577\|2020 | ···················· | ···················· | ···················· | ···················· | ···················· | ···················· |
| K | 6566 hCoV-19/England/MILK-9C40C3/2020\|EPI_ISL_566821\|20 | ···················· | ···················· | ···················· | ···················· | ···················· | ···················· |
| K | 6567 hCoV-19/England/MILK-9C3307/2020\|EPI_ISL_566822\|20 | ···················· | ···················· | ···················· | ···················· | ···················· | ···················· |
| K | 6568 hCoV-19/England/MILK-9C3079/2020\|EPI_ISL_566824\|20 | ···················· | ·A·················· | ···················· | ···················· | ···················· | ··A················· |
| K | 6569 hCoV-19/England/MILK-9C3AD5/2020\|EPI_ISL_566825\|20 | ···················· | ···················· | ···················· | ···················· | ···················· | ···················· |
| K | 6570 hCoV-19/England/MILK-9C345F/2020\|EPI_ISL_566826\|20 | ···················· | ·A·················· | ···················· | ···················· | ···················· | ··A················· |
| K | 6571 hCoV-19/England/MILK-9C4737/2020\|EPI_ISL_566827\|20 | ···················· | ···················· | ···················· | ···················· | ···················· | ···················· |
| K | 6572 hCoV-19/England/MILK-9C313A/2020\|EPI_ISL_566828\|20 | ···················· | ·A·················· | ···················· | ···················· | ···················· | ···················· |
| K | 6573 hCoV-19/England/MILK-9C403C/2020\|EPI_ISL_566829\|20 | ···················· | ···················· | ···················· | ···················· | ···················· | ···················· |
| K | 6574 hCoV-19/England/MILK-9C43AC/2020\|EPI_ISL_566830\|20 | ···················· | ···················· | ···················· | ···················· | ···················· | ···················· |
| K | 6575 hCoV-19/England/MILK-9C3F6D/2020\|EPI_ISL_566831\|20 | ···················· | ···················· | ···················· | ···················· | ···················· | ···················· |
| K | 6576 hCoV-19/England/MILK-9C349B/2020\|EPI_ISL_566832\|20 | ···················· | ···················· | ···················· | ···················· | ···················· | ···················· |
| K | 6577 hCoV-19/England/MILK-9C39BA/2020\|EPI_ISL_566833\|20 | ···················· | ···················· | ···················· | ···················· | ···················· | ···················· |

| | | | | | | | |
|---|---|---|---|---|---|---|---|
| K | 6578 hCoV-19/England/MILK-9C38DB/2020\|EPI_ISL_566834\|20 | ···················· | ···················· | ···················· | ···················· | ···················· | ···················· |
| K | 6579 hCoV-19/England/MILK-9C3CC0/2020\|EPI_ISL_566835\|20 | ···················· | ···················· | ···················· | ···················· | ···················· | ···················· |
| K | 6580 hCoV-19/England/MILK-9C3A6C/2020\|EPI_ISL_566836\|20 | ···················· | ·A·················· | ···················· | ···················· | ···················· | ···A················ |
| K | 6581 hCoV-19/England/MILK-9C411B/2020\|EPI_ISL_566837\|20 | ···················· | ···················· | ···················· | ···················· | ···················· | ···················· |
| K | 6582 hCoV-19/England/MILK-9C3ED9/2020\|EPI_ISL_566839\|20 | ···················· | ···················· | ···················· | ···················· | ···················· | ···················· |
| K | 6583 hCoV-19/England/MILK-9C33AD/2020\|EPI_ISL_566840\|20 | ···················· | ···················· | ···················· | ···················· | ···················· | ···················· |
| K | 6584 hCoV-19/England/MILK-9C3668/2020\|EPI_ISL_566842\|20 | ···················· | ·A·················· | ···················· | ···················· | ···················· | ···A················ |
| K | 6585 hCoV-19/England/MILK-9C3D09/2020\|EPI_ISL_566843\|20 | ···················· | ···················· | ···················· | ···················· | ···················· | ···················· |
| K | 6586 hCoV-19/England/MILK-9C41C0/2020\|EPI_ISL_566844\|20 | ···················· | ····A··············· | ···················· | ···················· | ···················· | ···················· |
| K | 6587 hCoV-19/England/MILK-9C3659/2020\|EPI_ISL_566845\|20 | ···················· | ···················· | ···················· | ···················· | ···················· | ···················· |
| K | 6588 hCoV-19/England/MILK-9C3EBB/2020\|EPI_ISL_566846\|20 | ···················· | ·A·················· | ···················· | ···················· | ···················· | ···A················ |
| K | 6589 hCoV-19/England/MILK-9C3A7B/2020\|EPI_ISL_566847\|20 | ···················· | ···················· | ···················· | ···················· | ···················· | ···················· |
| K | 6590 hCoV-19/England/MILK-9C41A2/2020\|EPI_ISL_566848\|20 | ···················· | ···················· | ···················· | ···················· | ···················· | ···················· |
| K | 6591 hCoV-19/England/MILK-9C3282/2020\|EPI_ISL_566849\|20 | ···················· | ·A·················· | ···················· | ···················· | ···················· | ···A················ |
| K | 6592 hCoV-19/England/MILK-9C437F/2020\|EPI_ISL_566850\|20 | ···················· | ···················· | ···················· | ···················· | ···················· | ···················· |
| K | 6593 hCoV-19/England/MILK-9C361D/2020\|EPI_ISL_566851\|20 | ···················· | ·A·················· | ···················· | ···················· | ···················· | ···A················ |
| K | 6594 hCoV-19/England/MILK-9C3B87/2020\|EPI_ISL_566852\|20 | ···················· | ···················· | ···················· | ···················· | ···················· | ···················· |
| K | 6596 hCoV-19/England/MILK-9C45D3/2020\|EPI_ISL_566854\|20 | ···················· | ···················· | ···················· | ···················· | ···················· | ···················· |
| K | 6597 hCoV-19/England/MILK-9C448B/2020\|EPI_ISL_566855\|20 | ···················· | ···················· | ···················· | ···················· | ···················· | ···················· |
| K | 6598 hCoV-19/England/MILK-9C3E24/2020\|EPI_ISL_566856\|20 | ···················· | ···················· | ···················· | ···················· | ···················· | ···················· |
| K | 6599 hCoV-19/England/MILK-9C4315/2020\|EPI_ISL_566857\|20 | ···················· | ···················· | ···················· | ···················· | ···················· | ···················· |
| K | 6600 hCoV-19/England/MILK-9C3167/2020\|EPI_ISL_566858\|20 | ···················· | ···················· | ···················· | ···················· | ···················· | ···················· |
| K | 6601 hCoV-19/England/MILK-9C31A3/2020\|EPI_ISL_566859\|20 | ···················· | ····A··············· | ···················· | ···················· | ···················· | ···················· |
| K | 6602 hCoV-19/England/MILK-9C40B4/2020\|EPI_ISL_566860\|20 | ···················· | ·A·················· | ···················· | ···················· | ···················· | ···A················ |
| K | 6603 hCoV-19/Wales/MILK-9C35F2/2020\|EPI_ISL_566862\|2020 | ···················· | ···················· | ···················· | ···················· | ···················· | ···················· |
| K | 6604 hCoV-19/Wales/MILK-9C42DC/2020\|EPI_ISL_566864\|2020 | ···················· | ···················· | ···················· | ···················· | ···················· | ···················· |
| K | 6605 hCoV-19/England/MILK-9C3DEB/2020\|EPI_ISL_566865\|20 | ···················· | ···················· | ···················· | ···················· | ···················· | ···················· |
| K | 6606 hCoV-19/England/MILK-9C3941/2020\|EPI_ISL_566867\|20 | ···················· | ·A·················· | ···················· | ···················· | ···················· | ···A················ |
| K | 6607 hCoV-19/Wales/MILK-9C39D8/2020\|EPI_ISL_566868\|2020 | ···················· | ···················· | ···················· | ···················· | ···················· | ···················· |
| K | 6608 hCoV-19/England/MILK-9C446D/2020\|EPI_ISL_566869\|20 | ···················· | ···················· | ···················· | ···················· | ···················· | ···················· |
| K | 6609 hCoV-19/England/MILK-9C42EB/2020\|EPI_ISL_566870\|20 | ···················· | ·A·················· | ···················· | ···················· | ···················· | ···A················ |
| K | 6610 hCoV-19/England/MILK-9C30C4/2020\|EPI_ISL_566871\|20 | ···················· | ···················· | ···················· | ···················· | ···················· | ···················· |
| K | 6611 hCoV-19/England/MILK-9C399C/2020\|EPI_ISL_566872\|20 | ···················· | ···················· | ···················· | ···················· | ······T············· | ···················· |
| K | 6612 hCoV-19/England/MILK-9C3EE8/2020\|EPI_ISL_566874\|20 | ···················· | ·A·················· | ···················· | ···················· | ···················· | ···A················ |
| K | 6613 hCoV-19/England/MILK-9C33DA/2020\|EPI_ISL_566875\|20 | ···················· | ···················· | ···················· | ···················· | ···················· | ···················· |
| K | 6614 hCoV-19/Wales/MILK-9C3D54/2020\|EPI_ISL_566877\|2020 | ···················· | ···················· | ···················· | ···················· | ···················· | ···················· |
| K | 6615 hCoV-19/England/MILK-9C3DDC/2020\|EPI_ISL_566879\|20 | ···················· | ···················· | ···················· | ···················· | ···················· | ···················· |
| K | 6616 hCoV-19/England/MILK-9C3D72/2020\|EPI_ISL_566880\|20 | ···················· | ···················· | ···················· | ···················· | ···················· | ···················· |
| K | 6617 hCoV-19/England/MILK-9C3431/2020\|EPI_ISL_566881\|20 | ···················· | ···················· | ···················· | ···················· | ···················· | ···················· |
| K | 6618 hCoV-19/England/MILK-9C3F03/2020\|EPI_ISL_566882\|20 | ···················· | ···················· | ···················· | ···················· | ···················· | ···················· |
| K | 6619 hCoV-19/England/MILK-9C4360/2020\|EPI_ISL_566883\|20 | ···················· | ···················· | ···················· | ···················· | ···················· | ···················· |
| K | 6620 hCoV-19/England/MILK-9C34B9/2020\|EPI_ISL_566884\|20 | ···················· | ·A·················· | ···················· | ···················· | ···················· | ···A················ |
| K | 6621 hCoV-19/England/MILK-9C3BB4/2020\|EPI_ISL_566885\|20 | ···················· | ···················· | ···················· | ···················· | ···················· | ···················· |
| K | 6622 hCoV-19/England/MILK-9C346E/2020\|EPI_ISL_566886\|20 | ···················· | ···················· | ···················· | ···················· | ···················· | ···················· |
| K | 6623 hCoV-19/Wales/MILK-9C3C48/2020\|EPI_ISL_566887\|2020 | ···················· | ···················· | ···················· | ···················· | ···················· | ···················· |
| K | 6624 hCoV-19/England/MILK-9C371A/2020\|EPI_ISL_566888\|20 | ···················· | ···················· | ···················· | ···················· | ···················· | ···················· |
| K | 6625 hCoV-19/England/MILK-9C4281/2020\|EPI_ISL_566889\|20 | ···················· | ···················· | ···················· | ···················· | ···················· | ···················· |
| K | 6626 hCoV-19/England/MILK-9C4306/2020\|EPI_ISL_566890\|20 | ···················· | ·A·················· | ···················· | ···················· | ···················· | ···A················ |
| K | 6627 hCoV-19/England/MILK-9C4342/2020\|EPI_ISL_566891\|20 | ···················· | ···················· | ···················· | ···················· | ···················· | ···················· |
| K | 6628 hCoV-19/England/MILK-9C4245/2020\|EPI_ISL_566892\|20 | ···················· | ···················· | ···················· | ···················· | ···················· | ···················· |
| K | 6629 hCoV-19/England/MILK-9C439D/2020\|EPI_ISL_566893\|20 | ···················· | ···················· | ···················· | ···················· | ···················· | ···················· |
| K | 6630 hCoV-19/England/MILK-9C3A02/2020\|EPI_ISL_566894\|20 | ···················· | ···················· | ···················· | ···················· | ···················· | ···················· |
| K | 6631 hCoV-19/England/MILK-9C312B/2020\|EPI_ISL_566895\|20 | ···················· | ···················· | ···················· | ···················· | ···················· | ···················· |
| K | 6632 hCoV-19/England/MILK-9C3BD2/2020\|EPI_ISL_566896\|20 | ···················· | ···················· | ···················· | ···················· | ···················· | ···················· |
| K | 6633 hCoV-19/England/MILK-9C3950/2020\|EPI_ISL_566898\|20 | ···················· | ···················· | ···················· | ···················· | ······T············· | ···················· |
| K | 6634 hCoV-19/England/MILK-9C3404/2020\|EPI_ISL_566899\|20 | ···················· | ···················· | ···················· | ···················· | ···················· | ···················· |
| K | 6635 hCoV-19/France/PAC-IHU-1011/2020\|EPI_ISL_569033\|20 | ···················· | ···················· | ···················· | ···················· | ···················· | ···················· |
| K | 6636 hCoV-19/France/PAC-IHU-1013/2020\|EPI_ISL_569035\|20 | ···················· | ····A··············· | ···················· | ···················· | ···················· | ···················· |
| K | 6637 hCoV-19/France/PAC-IHU-1014/2020\|EPI_ISL_569036\|20 | ···················· | ···················· | ···················· | ···················· | ···················· | ···················· |
| K | 6638 hCoV-19/France/PAC-IHU-1015/2020\|EPI_ISL_569037\|20 | ···················· | ····A··············· | ···················· | ···················· | ···················· | ···················· |
| K | 6639 hCoV-19/France/PAC-IHU-1018/2020\|EPI_ISL_569038\|20 | ···················· | ····A··············· | ···················· | ···················· | ···················· | ···················· |
| K | 6640 hCoV-19/Canada/ON-UHTC-0228/2020\|EPI_ISL_570011\|20 | ···················· | ···················· | ···················· | ···················· | ···················· | ···················· |
| K | 6641 hCoV-19/Canada/ON-UHTC-0239/2020\|EPI_ISL_570012\|20 | ···················· | ···················· | ···················· | ···················· | ···················· | ···················· |
| K | 6642 hCoV-19/Canada/ON-UHTC-0238/2020\|EPI_ISL_570019\|20 | ···················· | ···················· | ···················· | ···················· | ···················· | ···················· |
| K | 6643 hCoV-19/Canada/ON-UHTC-0230/2020\|EPI_ISL_570020\|20 | ···················· | ···················· | ···················· | ···················· | ···················· | ···················· |
| K | 6644 hCoV-19/Canada/ON-UHTC-0225/2020\|EPI_ISL_570021\|20 | ···················· | ···················· | ···················· | ···················· | ···················· | ···················· |
| K | 6645 hCoV-19/USA/ID-UW-6222/2020\|EPI_ISL_570088\|2020-04 | ···················· | ···················· | ···················· | ···················· | ···················· | ···················· |
| K | 6646 hCoV-19/USA/WA-UW-6238/2020\|EPI_ISL_570089\|2020-04 | ···················· | ···················· | ···················· | ···················· | ···················· | ···················· |
| K | 6647 hCoV-19/USA/WA-UW-6242/2020\|EPI_ISL_570090\|2020-04 | ···················· | ···················· | ···················· | ···················· | ···················· | ···················· |
| K | 6648 hCoV-19/USA/WA-UW-6244/2020\|EPI_ISL_570091\|2020-04 | ···················· | ···················· | ···················· | ···················· | ···················· | ···················· |
| K | 6649 hCoV-19/USA/CT-UW-6245/2020\|EPI_ISL_570092\|2020-04 | ···················· | ···················· | ···················· | ···················· | ···················· | ···················· |
| K | 6650 hCoV-19/USA/CT-UW-6260/2020\|EPI_ISL_570093\|2020-04 | ···················· | ···················· | ···················· | ···················· | ···················· | ···················· |
| K | 6651 hCoV-19/USA/CT-UW-6274/2020\|EPI_ISL_570094\|2020-04 | ···················· | ···················· | ···················· | ···················· | ···················· | ···················· |
| K | 6652 hCoV-19/USA/CT-UW-6288/2020\|EPI_ISL_570095\|2020-04 | ···················· | ···················· | ···················· | ···················· | ···················· | ···················· |
| K | 6653 hCoV-19/USA/WA-UW-6321/2020\|EPI_ISL_570096\|2020-04 | ···················· | ···················· | ···················· | ···················· | ···················· | ···················· |
| K | 6654 hCoV-19/USA/WA-UW-6339/2020\|EPI_ISL_570097\|2020-04 | ···················· | ···················· | ···················· | ···················· | ···················· | ···················· |
| K | 6655 hCoV-19/USA/WA-UW-6343/2020\|EPI_ISL_570098\|2020-04 | ···················· | ···················· | ···················· | ···················· | ···················· | ···················· |
| K | 6656 hCoV-19/USA/ID-UW-6348/2020\|EPI_ISL_570099\|2020-04 | ···················· | ···················· | ···················· | ···················· | ···················· | ···················· |
| K | 6657 hCoV-19/Switzerland/VD-ETHZ-280118/2020\|EPI_ISL_56 | ···················· | ···················· | ···················· | ···················· | ···················· | ···················· |
| K | 6658 hCoV-19/Switzerland/VD-ETHZ-280123/2020\|EPI_ISL_56 | ···················· | ···················· | ···················· | ···················· | ···················· | ···················· |

| | | | | | | | | | |
|---|---|---|---|---|---|---|---|---|---|
| K | 6659 | hCoV-19/Switzerland/VD-ETHZ-280125/2020\|EPI_ISL_56 | ···················· | ···················· | ···················· | ···················· | ···················· | ···················· | |
| K | 6660 | hCoV-19/Switzerland/VD-ETHZ-280129/2020\|EPI_ISL_56 | ···················· | ···················· | ···················· | ···················· | ···················· | ···················· | |
| K | 6661 | hCoV-19/Switzerland/VD-ETHZ-280136/2020\|EPI_ISL_56 | ···················· | ···················· | ···················· | ···················· | ···················· | ···················· | |
| K | 6662 | hCoV-19/Switzerland/VD-ETHZ-280143/2020\|EPI_ISL_56 | ···················· | ···················· | ···················· | ···················· | ···················· | ···················· | |
| K | 6663 | hCoV-19/Switzerland/VD-ETHZ-280144/2020\|EPI_ISL_56 | ···················· | ···················· | ···················· | ···················· | ···················· | ···················· | |
| K | 6664 | hCoV-19/Switzerland/VD-ETHZ-280145/2020\|EPI_ISL_56 | ···················· | ···················· | ···················· | ···················· | ···················· | ···················· | |
| K | 6665 | hCoV-19/Switzerland/VD-ETHZ-280146/2020\|EPI_ISL_56 | ···················· | ···················· | ···················· | ···················· | ···················· | ···················· | |
| K | 6666 | hCoV-19/Switzerland/VD-ETHZ-280147/2020\|EPI_ISL_56 | ···················· | ···················· | ···················· | ···················· | ···················· | ···················· | |
| K | 6667 | hCoV-19/Switzerland/VD-ETHZ-280158/2020\|EPI_ISL_56 | ···················· | ···················· | ···················· | ···················· | ···················· | ···················· | |
| K | 6668 | hCoV-19/Switzerland/VD-ETHZ-280160/2020\|EPI_ISL_56 | ···················· | ···················· | ···················· | ···················· | ···················· | ···················· | |
| K | 6669 | hCoV-19/Switzerland/VD-ETHZ-280163/2020\|EPI_ISL_56 | ···················· | ···················· | ···················· | ···················· | ···················· | ···················· | |
| K | 6670 | hCoV-19/Switzerland/VD-ETHZ-280164/2020\|EPI_ISL_56 | ···················· | ···················· | ···················· | ···················· | ···················· | ···················· | |
| K | 6671 | hCoV-19/Switzerland/VD-ETHZ-280165/2020\|EPI_ISL_56 | ···················· | ···················· | ···················· | ···················· | ···················· | ···················· | |
| K | 6672 | hCoV-19/Switzerland/VD-ETHZ-280166/2020\|EPI_ISL_56 | ···················· | ···················· | ···················· | ···················· | ···················· | ···················· | |
| K | 6673 | hCoV-19/Switzerland/VD-ETHZ-280167/2020\|EPI_ISL_56 | ···················· | ···················· | ···················· | ···················· | ···················· | ···················· | |
| K | 6674 | hCoV-19/Switzerland/VD-ETHZ-280168/2020\|EPI_ISL_56 | ···················· | ···················· | ···················· | ···················· | ···················· | ···················· | |
| K | 6675 | hCoV-19/Switzerland/VD-ETHZ-280170/2020\|EPI_ISL_56 | ···················· | ···················· | ···················· | ···················· | ···················· | ···················· | |
| K | 6676 | hCoV-19/Switzerland/VD-ETHZ-280181/2020\|EPI_ISL_56 | ···················· | ···················· | ···················· | ···················· | ···················· | ···················· | |
| K | 6677 | hCoV-19/Switzerland/VD-ETHZ-280202/2020\|EPI_ISL_56 | ···················· | ···················· | ···················· | ···················· | ···················· | ···················· | |
| K | 6678 | hCoV-19/Switzerland/VD-ETHZ-280203/2020\|EPI_ISL_56 | ···················· | ···················· | ···················· | ···················· | ···················· | ···················· | |
| K | 6679 | hCoV-19/Switzerland/VD-ETHZ-280205/2020\|EPI_ISL_56 | ···················· | ···················· | ···················· | ···················· | ···················· | ···················· | |
| K | 6680 | hCoV-19/Switzerland/VD-ETHZ-280206/2020\|EPI_ISL_56 | ···················· | ···················· | ···················· | ···················· | ···················· | ···················· | |
| K | 6682 | hCoV-19/Switzerland/VD-ETHZ-280222/2020\|EPI_ISL_56 | ···················· | ···················· | ···················· | ···················· | ···················· | ···················· | |
| K | 6683 | hCoV-19/Switzerland/VD-ETHZ-280229/2020\|EPI_ISL_56 | ···················· | ···················· | ···················· | ···················· | ···················· | ···················· | |
| K | 6684 | hCoV-19/Switzerland/VD-ETHZ-280230/2020\|EPI_ISL_56 | ···················· | ···················· | ···················· | ···················· | ···················· | ···················· | |
| K | 6685 | hCoV-19/Switzerland/VD-ETHZ-280231/2020\|EPI_ISL_56 | ···················· | ···················· | ···················· | ···················· | ···················· | ···················· | |
| K | 6686 | hCoV-19/Switzerland/VD-ETHZ-280238/2020\|EPI_ISL_56 | ···················· | ···················· | ···················· | ···················· | ···················· | ···················· | |
| K | 6687 | hCoV-19/Switzerland/VD-ETHZ-280240/2020\|EPI_ISL_56 | ···················· | ···················· | ···················· | ···················· | ···················· | ···················· | |
| K | 6688 | hCoV-19/Switzerland/VD-ETHZ-280241/2020\|EPI_ISL_56 | ···················· | ···················· | ···················· | ···················· | ···················· | ···················· | |
| K | 6689 | hCoV-19/Switzerland/VS-ETHZ-280128/2020\|EPI_ISL_56 | ···················· | ···················· | ···················· | ···················· | ···················· | ···················· | |
| K | 6690 | hCoV-19/Switzerland/VS-ETHZ-280204/2020\|EPI_ISL_56 | ···················· | ···················· | ···················· | ···················· | ···················· | ···················· | |
| K | 6691 | hCoV-19/Switzerland/VS-ETHZ-280210/2020\|EPI_ISL_56 | ···················· | ···················· | ···················· | ···················· | ···················· | ···················· | |
| K | 6692 | hCoV-19/Switzerland/VS-ETHZ-280218/2020\|EPI_ISL_56 | ···················· | ···················· | ···················· | ···················· | ···················· | ···················· | |
| K | 6693 | hCoV-19/Switzerland/VS-ETHZ-280237/2020\|EPI_ISL_56 | ···················· | ···················· | ···················· | ···················· | ···················· | ···················· | |
| K | 6694 | hCoV-19/Switzerland/ZH-ETHZ-280045/2020\|EPI_ISL_56 | ···················· | ···················· | ···················· | ···················· | ···················· | ···················· | |
| K | 6695 | hCoV-19/Switzerland/ZH-ETHZ-280073/2020\|EPI_ISL_56 | ···················· | ···················· | ···················· | ···················· | ···················· | ···················· | |
| K | 6696 | hCoV-19/Switzerland/ZH-ETHZ-280088/2020\|EPI_ISL_56 | ···················· | ···················· | ···················· | ···················· | ···················· | ···················· | |
| K | 6697 | hCoV-19/Switzerland/ZH-ETHZ-280119/2020\|EPI_ISL_56 | ···················· | ·A·················· | ···················· | ···················· | ···················· | ···················· | |
| K | 6698 | hCoV-19/Switzerland/ZH-ETHZ-280135/2020\|EPI_ISL_56 | ···················· | ···················· | ···················· | ···················· | ···················· | ···················· | |
| K | 6699 | hCoV-19/Switzerland/ZH-ETHZ-280169/2020\|EPI_ISL_56 | ···················· | ···················· | ···················· | ···················· | ···················· | ···················· | |
| K | 6700 | hCoV-19/Switzerland/ZH-ETHZ-280171/2020\|EPI_ISL_56 | ···················· | ···················· | ···················· | ···················· | ···················· | ···················· | |
| K | 6701 | hCoV-19/Switzerland/ZH-ETHZ-280172/2020\|EPI_ISL_56 | ···················· | ···················· | ···················· | ···················· | ···················· | ···················· | |
| K | 6702 | hCoV-19/Switzerland/ZH-ETHZ-280174/2020\|EPI_ISL_56 | ···················· | ···················· | ···················· | ···················· | ···················· | ···················· | |
| K | 6703 | hCoV-19/Switzerland/ZH-ETHZ-280196/2020\|EPI_ISL_56 | ···················· | ···················· | ···················· | ···················· | ···················· | ···················· | |
| K | 6704 | hCoV-19/Switzerland/ZH-ETHZ-280197/2020\|EPI_ISL_56 | ···················· | ···················· | ···················· | ···················· | ······K············· | ···················· | |
| K | 6705 | hCoV-19/Switzerland/ZH-ETHZ-280198/2020\|EPI_ISL_56 | ···················· | ···················· | ···················· | ···················· | ···················· | ···················· | |
| K | 6706 | hCoV-19/Switzerland/ZH-ETHZ-280209/2020\|EPI_ISL_56 | ···················· | ·A·················· | ···················· | ···················· | ···················· | ···················· | |
| K | 6707 | hCoV-19/USA/AK-PHL383/2020\|EPI_ISL_560554\|2020-05- | ···················· | ···················· | ···················· | ···················· | ···················· | ···················· | |
| K | 6708 | hCoV-19/USA/AK-PHL397/2020\|EPI_ISL_560555\|2020-06- | ···················· | ···················· | ···················· | ···················· | ···················· | ···················· | |
| K | 6709 | hCoV-19/USA/AK-PHL400/2020\|EPI_ISL_560556\|2020-06- | ···················· | ···················· | ···················· | ···················· | ···················· | ···················· | |
| K | 6710 | hCoV-19/USA/AK-PHL444/2020\|EPI_ISL_560557\|2020-06- | ···················· | ···················· | ···················· | ···················· | ···················· | ···················· | |
| K | 6711 | hCoV-19/USA/AK-PHL448/2020\|EPI_ISL_560558\|2020-06- | ···················· | ···················· | ···················· | ···················· | ···················· | ···················· | |
| K | 6712 | hCoV-19/USA/AK-PHL455/2020\|EPI_ISL_560559\|2020-06- | ···················· | ···················· | ···················· | ···················· | ···················· | ···················· | |
| K | 6713 | hCoV-19/USA/AK-PHL463/2020\|EPI_ISL_560560\|2020-06- | ···················· | ···················· | ···················· | ···················· | ···················· | ···················· | |
| K | 6714 | hCoV-19/USA/AK-PHL473/2020\|EPI_ISL_560561\|2020-06- | ···················· | ···················· | ···················· | ···················· | ···················· | ···················· | |
| K | 6715 | hCoV-19/England/GSTT-5446/2020\|EPI_ISL_560684\|2020 | ···················· | ···················· | ···················· | ···················· | ···················· | ···················· | |
| K | 6716 | hCoV-19/England/GSTT-5508/2020\|EPI_ISL_560686\|2020 | ···················· | ···················· | ···················· | ···················· | ···················· | ···················· | |
| K | 6717 | hCoV-19/Australia/VIC6714/2020\|EPI_ISL_564005\|2020 | ···················· | ···················· | ···················· | ···················· | ···················· | ···················· | |
| K | 6719 | hCoV-19/Australia/VIC6718/2020\|EPI_ISL_564009\|2020 | ···················· | ···················· | ···················· | ···················· | ···················· | ···················· | |
| K | 6720 | hCoV-19/Australia/VIC6727/2020\|EPI_ISL_564016\|2020 | ···················· | ···················· | ···················· | ···················· | ···················· | ···················· | |
| K | 6721 | hCoV-19/Australia/VIC6730/2020\|EPI_ISL_564017\|2020 | ···················· | ···················· | ···················· | ···················· | NNN················· | ···················· | 1 |
| K | 6722 | hCoV-19/Australia/VIC6731/2020\|EPI_ISL_564018\|2020 | ···················· | ···················· | ···················· | ···················· | ···················· | ···················· | |
| K | 6723 | hCoV-19/Australia/VIC6734/2020\|EPI_ISL_564019\|2020 | ···················· | ···················· | ···················· | ···················· | ···················· | ···················· | |
| K | 6724 | hCoV-19/Australia/VIC6735/2020\|EPI_ISL_564020\|2020 | ···················· | ···················· | ···················· | ···················· | ···················· | ···················· | |
| K | 6725 | hCoV-19/Australia/VIC6739/2020\|EPI_ISL_564021\|2020 | ···················· | ···················· | ···················· | ···················· | ···················· | ···················· | |
| K | 6726 | hCoV-19/Australia/VIC6740/2020\|EPI_ISL_564022\|2020 | ···················· | ···················· | ···················· | ···················· | ···················· | ···················· | |
| K | 6727 | hCoV-19/Australia/VIC6742/2020\|EPI_ISL_564023\|2020 | ···················· | ···················· | ···················· | ···················· | ···················· | ···················· | |
| K | 6728 | hCoV-19/Australia/VIC6744/2020\|EPI_ISL_564024\|2020 | ···················· | ···················· | ···················· | ···················· | ···················· | ···················· | |
| K | 6729 | hCoV-19/Australia/VIC6746/2020\|EPI_ISL_564025\|2020 | ···················· | ···················· | ···················· | ···················· | ···················· | ···················· | |
| K | 6730 | hCoV-19/Australia/VIC6747/2020\|EPI_ISL_564026\|2020 | ···················· | ···················· | ···················· | ···················· | ···················· | ···················· | |
| K | 6731 | hCoV-19/Australia/VIC6839/2020\|EPI_ISL_564090\|2020 | ···················· | ···················· | ···················· | ···················· | ···················· | ···················· | |
| K | 6732 | hCoV-19/Australia/VIC6844/2020\|EPI_ISL_564091\|2020 | ···················· | ···················· | ···················· | ···················· | ···················· | ···················· | |
| K | 6733 | hCoV-19/Australia/VIC6845/2020\|EPI_ISL_564092\|2020 | ···················· | ···················· | ···················· | ···················· | ···················· | ···················· | |
| K | 6734 | hCoV-19/Australia/VIC6846/2020\|EPI_ISL_564093\|2020 | ···················· | ···················· | ···················· | ···················· | ···················· | ···················· | |
| K | 6735 | hCoV-19/Australia/VIC6847/2020\|EPI_ISL_564094\|2020 | ···················· | ···················· | ···················· | ···················· | ···················· | ···················· | |
| K | 6736 | hCoV-19/Australia/VIC6850/2020\|EPI_ISL_564095\|2020 | ···················· | ···················· | ···················· | ···················· | ···················· | ···················· | |
| K | 6737 | hCoV-19/Australia/VIC6851/2020\|EPI_ISL_564096\|2020 | ···················· | ···················· | ···················· | ···················· | ···················· | ···················· | |
| K | 6738 | hCoV-19/Australia/VIC6853/2020\|EPI_ISL_564097\|2020 | ···················· | ···················· | ···················· | ···················· | ···················· | ···················· | |
| K | 6739 | hCoV-19/Australia/VIC6854/2020\|EPI_ISL_564098\|2020 | ···················· | ···················· | ···················· | ···················· | ···················· | ···················· | |
| K | 6740 | hCoV-19/Australia/VIC6856/2020\|EPI_ISL_564099\|2020 | ···················· | ···················· | ···················· | ···················· | ···················· | ···················· | |

| | | | | | | | |
|---|---|---|---|---|---|---|---|
| K | 6741 hCoV-19/Australia/VIC8986/2020\|EPI_ISL_565336\|2020 | ···················· | ···················· | ···················· | ···················· | ···················· | ···················· |
| K | 6742 hCoV-19/Australia/VIC8987/2020\|EPI_ISL_565337\|2020 | ···················· | ······A············· | ···················· | ···················· | ···················· | ···················· |
| K | 6743 hCoV-19/Australia/VIC8997/2020\|EPI_ISL_565347\|2020 | ···················· | ······A············· | ···················· | ···················· | ···················· | ···················· |
| K | 6744 hCoV-19/Australia/VIC8998/2020\|EPI_ISL_565348\|2020 | ···················· | ······A············· | ···················· | ···················· | ···················· | ···················· |
| K | 6745 hCoV-19/Australia/VIC8999/2020\|EPI_ISL_565349\|2020 | ···················· | ···················· | ···················· | ···················· | ···················· | ···················· |
| K | 6746 hCoV-19/Australia/VIC9000/2020\|EPI_ISL_565350\|2020 | ···················· | ···················· | ···················· | ···················· | ···················· | ···················· |
| K | 6747 hCoV-19/Australia/VIC9007/2020\|EPI_ISL_565351\|2020 | ···················· | ···················· | ···················· | ···················· | ···················· | ···················· |
| K | 6748 hCoV-19/Australia/VIC9008/2020\|EPI_ISL_565352\|2020 | ···················· | ···················· | ···················· | ···················· | ···················· | ···················· |
| K | 6750 hCoV-19/Australia/VIC9011/2020\|EPI_ISL_565354\|2020 | ···················· | ···················· | ···················· | ···················· | ···················· | ···················· |
| K | 6751 hCoV-19/Australia/VIC9018/2020\|EPI_ISL_565355\|2020 | ···················· | ······A············· | ···················· | ···················· | ···················· | ···················· |
| K | 6752 hCoV-19/Australia/VIC9019/2020\|EPI_ISL_565356\|2020 | ···················· | ···················· | ···················· | ···················· | ···················· | ···················· |
| K | 6753 hCoV-19/Australia/VIC9020/2020\|EPI_ISL_565357\|2020 | ···················· | ···················· | ···················· | ···················· | ···················· | ···················· |
| K | 6754 hCoV-19/England/MILK-9C2D55/2020\|EPI_ISL_566668\|20 | ···················· | ···················· | ···················· | ···················· | ···················· | ···················· |
| K | 6755 hCoV-19/England/MILK-9C2C76/2020\|EPI_ISL_566678\|20 | ···················· | ···················· | ···················· | ···················· | ···················· | ···················· |
| K | 6756 hCoV-19/England/MILK-9C31EF/2020\|EPI_ISL_566679\|20 | ···················· | ···················· | ···················· | ···················· | ···················· | ···················· |
| K | 6757 hCoV-19/England/MILK-9C410C/2020\|EPI_ISL_566680\|20 | ···················· | ···················· | ···················· | ···················· | ···················· | ···················· |
| K | 6758 hCoV-19/England/MILK-9C3A5D/2020\|EPI_ISL_566681\|20 | ···················· | ···················· | ···················· | ···················· | ···················· | ···················· |
| K | 6759 hCoV-19/England/MILK-9C438E/2020\|EPI_ISL_566682\|20 | ···················· | ···················· | ···················· | ···················· | ···················· | ···················· |
| K | 6760 hCoV-19/Wales/MILK-9C3AB7/2020\|EPI_ISL_566683\|2020 | ···················· | ···················· | ···················· | ···················· | ···················· | ···················· |
| K | 6761 hCoV-19/England/MILK-9C3677/2020\|EPI_ISL_566684\|20 | ···················· | ·A·················· | ···················· | ···················· | ···················· | ···A················ |
| K | 6762 hCoV-19/England/MILK-9C3D18/2020\|EPI_ISL_566685\|20 | ···················· | ···················· | ···················· | ···················· | ···················· | ···················· |
| K | 6763 hCoV-19/Wales/MILK-9C2F9B/2020\|EPI_ISL_566686\|2020 | ···················· | ···················· | ···················· | ···················· | ···················· | ···················· |
| K | 6764 hCoV-19/England/MILK-9C3CFD/2020\|EPI_ISL_566687\|20 | ···················· | ·A·······A·········· | ···················· | ···················· | ···················· | ···················· |
| K | 6765 hCoV-19/England/MILK-9C3F7C/2020\|EPI_ISL_566688\|20 | ···················· | ···················· | ···················· | ···················· | ···················· | ···················· |
| K | 6766 hCoV-19/England/MILK-9C4403/2020\|EPI_ISL_566801\|20 | ···················· | ···················· | ···················· | ···················· | ···················· | ···················· |
| K | 6767 hCoV-19/England/MILK-9C3501/2020\|EPI_ISL_566802\|20 | ···················· | ···················· | ···················· | ···················· | ···················· | ···················· |
| K | 6768 hCoV-19/England/MILK-9C4069/2020\|EPI_ISL_566803\|20 | ···················· | ···················· | ···················· | ···················· | ···················· | ···················· |
| K | 6769 hCoV-19/England/MILK-9C3334/2020\|EPI_ISL_566804\|20 | ···················· | ···················· | ···················· | ···················· | ···················· | ···················· |
| K | 6770 hCoV-19/England/MILK-9C34D7/2020\|EPI_ISL_566806\|20 | ···················· | ···················· | ···················· | ···················· | ···················· | ···················· |
| K | 6771 hCoV-19/England/MILK-9C45B5/2020\|EPI_ISL_566807\|20 | ···················· | ···················· | ···················· | ···················· | ···················· | ···················· |
| K | 6772 hCoV-19/Wales/MILK-9C353E/2020\|EPI_ISL_566808\|2020 | ···················· | ···················· | ···················· | ···················· | ···················· | ···················· |
| K | 6773 hCoV-19/England/MILK-9C3D63/2020\|EPI_ISL_566809\|20 | ···················· | ···················· | ···················· | ···················· | ···················· | ···················· |
| K | 6774 hCoV-19/England/MILK-9C356B/2020\|EPI_ISL_566810\|20 | ···················· | ···················· | ···················· | ···················· | ···················· | ···················· |
| K | 6775 hCoV-19/England/MILK-9C363B/2020\|EPI_ISL_566812\|20 | ···················· | ···················· | ···················· | ···················· | ···················· | ···················· |
| K | 6776 hCoV-19/England/MILK-9C46C1/2020\|EPI_ISL_566813\|20 | ···················· | ···················· | ···················· | ···················· | ···················· | ···················· |
| K | 6777 hCoV-19/England/MILK-9C3CA2/2020\|EPI_ISL_566814\|20 | ···················· | ···················· | ···················· | ···················· | ···················· | ···················· |
| K | 6778 hCoV-19/England/MILK-9C32CE/2020\|EPI_ISL_566816\|20 | ···················· | ·A·················· | ···················· | ···················· | ···················· | ···A················ |
| K | 6779 hCoV-19/England/MILK-9C3291/2020\|EPI_ISL_566817\|20 | ···················· | ···················· | ···················· | ···················· | ···················· | ···················· |
| K | 6780 hCoV-19/England/MILK-9C43BB/2020\|EPI_ISL_566818\|20 | ···················· | ···················· | ···················· | ···················· | ···················· | ···················· |
| K | 6781 hCoV-19/England/MILK-9C3AA8/2020\|EPI_ISL_566819\|20 | ···················· | ···················· | ···················· | ···················· | ···················· | ···················· |
| K | 6782 hCoV-19/England/MILK-9C3AE4/2020\|EPI_ISL_566820\|20 | ···················· | ···················· | ···················· | ···················· | ······T············· | ···················· |
| K | 6783 hCoV-19/Wales/ALDP-9BE475/2020\|EPI_ISL_567998\|2020 | ···················· | ···················· | ···················· | ···················· | ···················· | ···················· |
| K | 6784 hCoV-19/France/PAC-IHU-0973/2020\|EPI_ISL_569020\|20 | ···················· | ···················· | ···················· | ···················· | ···················· | ···················· |
| K | 6785 hCoV-19/France/PAC-IHU-0974/2020\|EPI_ISL_569021\|20 | ···················· | ···················· | ···················· | ···················· | ···················· | ···················· |
| K | 6786 hCoV-19/France/PAC-IHU-0975/2020\|EPI_ISL_569022\|20 | ···················· | ···················· | ···················· | ···················· | ···················· | ···················· |
| K | 6787 hCoV-19/France/PAC-IHU-0976/2020\|EPI_ISL_569023\|20 | ···················· | ····A··············· | ···················· | ···················· | ···················· | ···················· |
| K | 6788 hCoV-19/France/PAC-IHU-0977/2020\|EPI_ISL_569024\|20 | ···················· | ···················· | ···················· | ···················· | ···················· | ···················· |
| K | 6789 hCoV-19/France/PAC-IHU-0978/2020\|EPI_ISL_569025\|20 | ···················· | ····A··············· | ···················· | ···················· | ···················· | ···················· |
| K | 6790 hCoV-19/France/PAC-IHU-0979/2020\|EPI_ISL_569026\|20 | ···················· | ····A··············· | ···················· | ···················· | ···················· | ···················· |
| K | 6791 hCoV-19/France/PAC-IHU-0980/2020\|EPI_ISL_569027\|20 | ···················· | ····A··············· | ···················· | ···················· | ···················· | ···················· |
| K | 6792 hCoV-19/France/PAC-IHU-0981/2020\|EPI_ISL_569028\|20 | ···················· | ·A·················· | ···················· | ···················· | ···················· | ···················· |
| K | 6793 hCoV-19/France/PAC-IHU-0982/2020\|EPI_ISL_569029\|20 | ···················· | ····A··············· | ···················· | ···················· | ···················· | ···················· |
| K | 6794 hCoV-19/France/PAC-IHU-0985/2020\|EPI_ISL_569031\|20 | ···················· | ···················· | ···················· | ···················· | ···················· | ···················· |
| K | 6795 hCoV-19/France/PAC-IHU-0986/2020\|EPI_ISL_569032\|20 | ···················· | ···················· | ···················· | ···················· | ···················· | ···················· |
| K | 6796 hCoV-19/France/PAC-IHU-1012/2020\|EPI_ISL_569034\|20 | ···················· | ····A··············· | ···················· | ···················· | ···················· | ···················· |
| K | 6797 hCoV-19/France/PAC-IHU-1054/2020\|EPI_ISL_569041\|20 | ···················· | ···················· | ···················· | ···················· | ···················· | ···················· |
| K | 6798 hCoV-19/France/PAC-IHU-1058/2020\|EPI_ISL_569043\|20 | ···················· | ···················· | ···················· | ···················· | ···················· | ···················· |
| K | 6799 hCoV-19/France/PAC-IHU-1064/2020\|EPI_ISL_569044\|20 | ···················· | ···················· | ···················· | ···················· | ···················· | ···················· |
| K | 6800 hCoV-19/France/PAC-IHU-1065/2020\|EPI_ISL_569045\|20 | ···················· | ···················· | ···················· | ···················· | ···················· | ···················· |
| K | 6801 hCoV-19/France/PAC-IHU-1068/2020\|EPI_ISL_569046\|20 | ···················· | ···················· | ···················· | ···················· | ···················· | ···················· |
| K | 6802 hCoV-19/France/PAC-IHU-1085/2020\|EPI_ISL_569047\|20 | ···················· | ·A·················· | ···················· | ···················· | ···················· | ···················· |
| K | 6803 hCoV-19/France/PAC-IHU-1086/2020\|EPI_ISL_569048\|20 | ···················· | ···················· | ···················· | ···················· | ···················· | ···················· |
| K | 6804 hCoV-19/France/PAC-IHU-1087/2020\|EPI_ISL_569049\|20 | ···················· | ·A·················· | ···················· | ···················· | ···················· | ···················· |
| K | 6805 hCoV-19/France/PAC-IHU-1092/2020\|EPI_ISL_569050\|20 | ···················· | ···················· | ···················· | ···················· | ···················· | ···················· |
| K | 6806 hCoV-19/France/PAC-IHU-1097/2020\|EPI_ISL_569051\|20 | ···················· | ···················· | ···················· | ···················· | ···················· | ···················· |
| K | 6807 hCoV-19/France/PAC-IHU-1126/2020\|EPI_ISL_569053\|20 | ···················· | ···················· | ···················· | ···················· | ···················· | ···················· |
| K | 6808 hCoV-19/France/PAC-IHU-1128/2020\|EPI_ISL_569054\|20 | ···················· | ···················· | ···················· | ···················· | ···················· | ···················· |
| K | 6809 hCoV-19/France/PAC-IHU-1131/2020\|EPI_ISL_569055\|20 | ···················· | ···················· | ···················· | ···················· | ···················· | ···················· |
| K | 6810 hCoV-19/France/PAC-IHU-1134/2020\|EPI_ISL_569056\|20 | ···················· | ···················· | ···················· | ···················· | ···················· | ···················· |
| K | 6811 hCoV-19/France/PAC-IHU-1135/2020\|EPI_ISL_569057\|20 | ···················· | ···················· | ···················· | ···················· | ···················· | ···················· |
| K | 6812 hCoV-19/France/PAC-IHU-1138/2020\|EPI_ISL_569058\|20 | ···················· | ···················· | ···················· | ···················· | ···················· | ···················· |
| K | 6813 hCoV-19/France/PAC-IHU-1146/2020\|EPI_ISL_569060\|20 | ···················· | ···················· | ···················· | ···················· | ···················· | ···················· |
| K | 6814 hCoV-19/France/PAC-IHU-1151/2020\|EPI_ISL_569061\|20 | ···················· | ·A·················· | ···················· | ···················· | ···················· | ···················· |
| K | 6815 hCoV-19/France/PAC-IHU-1155/2020\|EPI_ISL_569062\|20 | ···················· | ···················· | ···················· | ···················· | ···················· | ···················· |
| K | 6816 hCoV-19/France/PAC-IHU-1157/2020\|EPI_ISL_569063\|20 | ···················· | ···················· | ···················· | ···················· | ···················· | ···················· |
| K | 6817 hCoV-19/France/PAC-IHU-1158/2020\|EPI_ISL_569064\|20 | ···················· | ·A·················· | ···················· | ···················· | ···················· | ···················· |
| K | 6818 hCoV-19/France/PAC-IHU-1162/2020\|EPI_ISL_569067\|20 | ···················· | ····A··············· | ···················· | ···················· | ···················· | ···················· |
| K | 6819 hCoV-19/France/PAC-IHU-1164/2020\|EPI_ISL_569068\|20 | ···················· | ···················· | ···················· | ···················· | ···················· | ···················· |
| K | 6820 hCoV-19/France/PAC-IHU-1166/2020\|EPI_ISL_569070\|20 | ···················· | ···················· | ···················· | ···················· | ···················· | ···················· |
| K | 6821 hCoV-19/France/PAC-IHU-1167/2020\|EPI_ISL_569071\|20 | ···················· | ···················· | ···················· | ···················· | ···················· | ···················· |

| | | | | | | | |
|---|---|---|---|---|---|---|---|
| K | 6822 hCoV-19/France/PAC-IHU-1169/2020\|EPI_ISL_569072\|20 | ···················· | ···················· | ···················· | ···················· | ···················· | ···················· |
| K | 6823 hCoV-19/France/PAC-IHU-1171/2020\|EPI_ISL_569073\|20 | ···················· | ···················· | ···················· | ···················· | ···················· | ···················· |
| K | 6824 hCoV-19/France/PAC-IHU-1172/2020\|EPI_ISL_569074\|20 | ···················· | ···················· | ···················· | ···················· | ···················· | ···················· |
| K | 6825 hCoV-19/France/PAC-IHU-1173/2020\|EPI_ISL_569075\|20 | ···················· | ···················· | ···················· | ···················· | ···················· | ···················· |
| K | 6826 hCoV-19/France/PAC-IHU-1175/2020\|EPI_ISL_569076\|20 | ···················· | ·A·················· | ···················· | ···················· | ···················· | ···················· |
| K | 6827 hCoV-19/France/PAC-IHU-1176/2020\|EPI_ISL_569077\|20 | ···················· | ···················· | ···················· | ···················· | ···················· | ···················· |
| K | 6828 hCoV-19/France/PAC-IHU-1179/2020\|EPI_ISL_569078\|20 | ···················· | ···················· | ···················· | ···················· | ···················· | ···················· |
| K | 6829 hCoV-19/France/PAC-IHU-1189/2020\|EPI_ISL_569079\|20 | ···················· | ···················· | ···················· | ···················· | ···················· | ···················· |
| K | 6830 hCoV-19/France/PAC-IHU-1191/2020\|EPI_ISL_569080\|20 | ···················· | ···················· | ···················· | ···················· | ···················· | ···················· |
| K | 6831 hCoV-19/France/PAC-IHU-1193/2020\|EPI_ISL_569081\|20 | ···················· | ·A·················· | ···················· | ···················· | ···················· | ···················· |
| K | 6832 hCoV-19/France/PAC-IHU-1195/2020\|EPI_ISL_569082\|20 | ···················· | ···················· | ···················· | ···················· | ···················· | ···················· |
| K | 6833 hCoV-19/France/PAC-IHU-1206/2020\|EPI_ISL_569084\|20 | ···················· | ···················· | ···················· | ···················· | ···················· | ···················· |
| K | 6834 hCoV-19/France/PAC-IHU-1207/2020\|EPI_ISL_569085\|20 | ···················· | ···················· | ···················· | ···················· | ···················· | ···················· |
| K | 6835 hCoV-19/France/PAC-IHU-1210/2020\|EPI_ISL_569086\|20 | ···················· | ···················· | ···················· | ···················· | ···················· | ···················· |
| K | 6836 hCoV-19/France/PAC-IHU-1211/2020\|EPI_ISL_569087\|20 | ···················· | ···················· | ···················· | ···················· | ···················· | ···················· |
| K | 6837 hCoV-19/France/PAC-IHU-1212/2020\|EPI_ISL_569088\|20 | ···················· | ···A················ | ···················· | ···················· | ···················· | ···················· |
| K | 6838 hCoV-19/France/PAC-IHU-1213/2020\|EPI_ISL_569089\|20 | ···················· | ···A················ | ···················· | ···················· | ···················· | ···················· |
| K | 6839 hCoV-19/France/PAC-IHU-1215/2020\|EPI_ISL_569090\|20 | ···················· | ···A················ | ···················· | ···················· | ···················· | ···················· |
| K | 6840 hCoV-19/France/PAC-IHU-1223/2020\|EPI_ISL_569091\|20 | ···················· | ···················· | ···················· | ···················· | ···················· | ···················· |
| K | 6841 hCoV-19/France/PAC-IHU-1224/2020\|EPI_ISL_569092\|20 | ···················· | ···················· | ···················· | ···················· | ···················· | ···················· |
| K | 6842 hCoV-19/France/PAC-IHU-1229/2020\|EPI_ISL_569093\|20 | ···················· | ·A·················· | ···················· | ···················· | ···················· | ···················· |
| K | 6843 hCoV-19/France/PAC-IHU-1230/2020\|EPI_ISL_569094\|20 | ···················· | ···A················ | ···················· | ···················· | ···················· | ···················· |
| K | 6844 hCoV-19/France/PAC-IHU-1250/2020\|EPI_ISL_569096\|20 | ···················· | ···················· | ···················· | ···················· | ···················· | ···················· |
| K | 6845 hCoV-19/France/PAC-IHU-1251/2020\|EPI_ISL_569097\|20 | ···················· | ···················· | ···················· | ···················· | ···················· | ···················· |
| K | 6846 hCoV-19/France/PAC-IHU-1255/2020\|EPI_ISL_569098\|20 | ···················· | ···················· | ···················· | ···················· | ···················· | ···················· |
| K | 6847 hCoV-19/France/PAC-IHU-1258/2020\|EPI_ISL_569099\|20 | ···················· | ·A·················· | ···················· | ···················· | ···················· | ···················· |
| K | 6848 hCoV-19/Canada/ON-UHTC-0271/2020\|EPI_ISL_570000\|20 | ···················· | ···················· | ···················· | ···················· | ···················· | ···················· |
| K | 6849 hCoV-19/Canada/ON-UHTC-0261/2020\|EPI_ISL_570001\|20 | ···················· | ···················· | ···················· | ···················· | ···················· | ···················· |
| K | 6850 hCoV-19/Canada/ON-UHTC-0270/2020\|EPI_ISL_570002\|20 | ···················· | ·A·················· | ···················· | ···················· | ···················· | ···A················ |
| K | 6851 hCoV-19/Canada/ON-UHTC-0277/2020\|EPI_ISL_570003\|20 | ···················· | ···················· | ···················· | ···················· | ···················· | ···················· |
| K | 6852 hCoV-19/Canada/ON-UHTC-0257/2020\|EPI_ISL_570004\|20 | ···················· | ···················· | ···················· | ···················· | ···················· | ···················· |
| K | 6853 hCoV-19/Canada/ON-UHTC-0275/2020\|EPI_ISL_570005\|20 | ···················· | ···················· | ···················· | ···················· | ···················· | ···················· |
| K | 6854 hCoV-19/Canada/ON-UHTC-0262/2020\|EPI_ISL_570006\|20 | ···················· | ···················· | ···················· | ···················· | ···················· | ···················· |
| K | 6855 hCoV-19/Canada/ON-UHTC-0253/2020\|EPI_ISL_570007\|20 | ···················· | ···················· | ···················· | ···················· | ···················· | ···················· |
| K | 6856 hCoV-19/Canada/ON-UHTC-0278/2020\|EPI_ISL_570008\|20 | ···················· | ···················· | ···················· | ···················· | ···················· | ···················· |
| K | 6857 hCoV-19/Canada/ON-UHTC-0241/2020\|EPI_ISL_570009\|20 | ···················· | ···················· | ···················· | ···················· | ···················· | ···················· |
| K | 6858 hCoV-19/Canada/ON-UHTC-0240/2020\|EPI_ISL_570014\|20 | ···················· | ···················· | ···················· | ···················· | ···················· | ···················· |
| K | 6859 hCoV-19/Canada/ON-UHTC-0243/2020\|EPI_ISL_570016\|20 | ···················· | ···················· | ···················· | ···················· | ···················· | ···················· |
| K | 6860 hCoV-19/Canada/ON-UHTC-0222/2020\|EPI_ISL_570017\|20 | ···················· | ···················· | ···················· | ···················· | ···················· | ···················· |
| K | 6861 hCoV-19/Canada/ON-UHTC-0218/2020\|EPI_ISL_570023\|20 | ···················· | ···················· | ···················· | ···················· | ···················· | ···················· |
| K | 6862 hCoV-19/Canada/ON-UHTC-0245/2020\|EPI_ISL_570025\|20 | ···················· | ···················· | ···················· | ···················· | ···················· | ···················· |
| K | 6863 hCoV-19/USA/WA-UW-335/2020\|EPI_ISL_570028\|2020-03- | ···················· | ···················· | ···················· | ···················· | ···················· | ···················· |
| K | 6864 hCoV-19/USA/WA-UW-3560/2020\|EPI_ISL_570029\|2020-03 | ···················· | ···················· | ···················· | ···················· | ···················· | ···················· |
| K | 6865 hCoV-19/USA/WA-UW-3562/2020\|EPI_ISL_570030\|2020-03 | ···················· | ···················· | ···················· | ···················· | ···················· | ···················· |
| K | 6866 hCoV-19/USA/WA-UW-3584/2020\|EPI_ISL_570032\|2020-03 | ···················· | ···················· | ···················· | ···················· | ···················· | ···················· |
| K | 6867 hCoV-19/USA/WA-UW-3600/2020\|EPI_ISL_570033\|2020-03 | ···················· | ···················· | ···················· | ···················· | ···················· | ···················· |
| K | 6868 hCoV-19/USA/WA-UW-3607/2020\|EPI_ISL_570034\|2020-03 | ···················· | ···················· | ···················· | ···················· | ···················· | ···················· |
| K | 6869 hCoV-19/USA/WA-UW-3616/2020\|EPI_ISL_570037\|2020-04 | ···················· | ···················· | ···················· | A··················· | ···················· | ···················· |
| K | 6870 hCoV-19/USA/WA-UW-3630/2020\|EPI_ISL_570038\|2020-04 | ···················· | ···················· | ···················· | ···················· | ···················· | ···················· |
| K | 6871 hCoV-19/USA/WA-UW-3634/2020\|EPI_ISL_570040\|2020-03 | ···················· | ···················· | ···················· | ···················· | ···················· | ···················· |
| K | 6872 hCoV-19/USA/WA-UW-3662/2020\|EPI_ISL_570041\|2020-03 | ···················· | ···················· | ···················· | ···················· | ···················· | ···················· |
| K | 6873 hCoV-19/USA/CT-UW-3692/2020\|EPI_ISL_570042\|2020-03 | ···················· | ···················· | ···················· | ···················· | ···················· | ···················· |
| K | 6874 hCoV-19/USA/WA-UW-3783/2020\|EPI_ISL_570043\|2020-04 | ···················· | ···················· | ···················· | ···················· | ···················· | ···················· |
| K | 6875 hCoV-19/USA/WA-UW-383/2020\|EPI_ISL_570044\|2020-03- | ···················· | ···················· | ···················· | ···················· | ···················· | ···················· |
| K | 6876 hCoV-19/USA/WA-UW-4303/2020\|EPI_ISL_570046\|2020-03 | ···················· | ···················· | ···················· | ···················· | ···················· | ···················· |
| K | 6877 hCoV-19/USA/WA-UW-4331/2020\|EPI_ISL_570047\|2020-03 | ···················· | ···················· | ···················· | ···················· | ···················· | ···················· |
| K | 6878 hCoV-19/USA/ID-UW-4417/2020\|EPI_ISL_570048\|2020-03 | ···················· | ···················· | ···················· | ···················· | ···················· | ···················· |
| K | 6879 hCoV-19/USA/WA-UW-4434/2020\|EPI_ISL_570049\|2020-05 | ···················· | ···················· | ···················· | ···················· | ···················· | ···················· |
| K | 6880 hCoV-19/USA/WA-UW-4541/2020\|EPI_ISL_570050\|2020-03 | ···················· | ···················· | ···················· | ···················· | ···················· | ···················· |
| K | 6881 hCoV-19/USA/WA-UW-4583/2020\|EPI_ISL_570051\|2020-03 | ···················· | ···················· | ···················· | ···················· | ···················· | ···················· |
| K | 6882 hCoV-19/USA/WA-UW-4630/2020\|EPI_ISL_570052\|2020-03 | ···················· | ···················· | ···················· | ···················· | ···················· | ···················· |
| K | 6883 hCoV-19/USA/WA-UW-4640/2020\|EPI_ISL_570053\|2020-03 | ···················· | ···················· | ···················· | ···················· | ···················· | ···················· |
| K | 6884 hCoV-19/USA/WA-UW-4725/2020\|EPI_ISL_570054\|2020-04 | ···················· | ···················· | ···················· | ···················· | ···················· | ···················· |
| K | 6885 hCoV-19/USA/WA-UW-4727/2020\|EPI_ISL_570056\|2020-04 | ···················· | ···················· | ···················· | ···················· | ···················· | ···················· |
| K | 6886 hCoV-19/USA/WA-UW-4730/2020\|EPI_ISL_570057\|2020-04 | ···················· | ···················· | ···················· | ···················· | ···················· | ···················· |
| K | 6887 hCoV-19/USA/WA-UW-4748/2020\|EPI_ISL_570059\|2020-03 | ···················· | ···················· | ···················· | ···················· | ···················· | ···················· |
| K | 6888 hCoV-19/USA/WA-UW-4759/2020\|EPI_ISL_570060\|2020-03 | ···················· | ···················· | ···················· | ···················· | ···················· | ···················· |
| K | 6889 hCoV-19/USA/WA-UW-5161/2020\|EPI_ISL_570061\|2020-04 | ···················· | ···················· | ···················· | ···················· | ···················· | ···················· |
| K | 6890 hCoV-19/USA/WA-UW-520/2020\|EPI_ISL_570062\|2020-03- | ···················· | ···················· | ···················· | ···················· | ···················· | ···················· |
| K | 6891 hCoV-19/USA/WA-UW-5201/2020\|EPI_ISL_570063\|2020-04 | ···················· | ···················· | ···················· | ···················· | ···················· | ···················· |
| K | 6892 hCoV-19/USA/WA-UW-523/2020\|EPI_ISL_570064\|2020-03- | ···················· | ···················· | ···················· | ···················· | ···················· | ···················· |
| K | 6893 hCoV-19/USA/WA-UW-529/2020\|EPI_ISL_570065\|2020-03- | ···················· | ···················· | ···················· | ···················· | ···················· | ···················· |
| K | 6894 hCoV-19/USA/WA-UW-5369/2020\|EPI_ISL_570066\|2020-04 | ···················· | ···················· | ···················· | ···················· | ···················· | ···················· |
| K | 6895 hCoV-19/USA/WA-UW-5382/2020\|EPI_ISL_570067\|2020-04 | ···················· | ···················· | ···················· | ···················· | ···················· | ···················· |
| K | 6896 hCoV-19/USA/WA-UW-5403/2020\|EPI_ISL_570068\|2020-03 | ···················· | ···················· | ···················· | ···················· | ···················· | ···················· |
| K | 6897 hCoV-19/USA/WA-UW-5428/2020\|EPI_ISL_570069\|2020-03 | ···················· | ···················· | ···················· | ···················· | ···················· | ···················· |
| K | 6898 hCoV-19/USA/WA-UW-5606/2020\|EPI_ISL_570071\|2020-04 | ···················· | ···················· | ···················· | ···················· | ···················· | ···················· |
| K | 6899 hCoV-19/USA/WA-UW-5670/2020\|EPI_ISL_570072\|2020-04 | ···················· | ···················· | ···················· | ···················· | ···················· | ···················· |
| K | 6900 hCoV-19/USA/WA-UW-5731/2020\|EPI_ISL_570073\|2020-04 | ···················· | ···················· | ···················· | ···················· | ···················· | ···················· |
| K | 6901 hCoV-19/USA/WA-UW-5746/2020\|EPI_ISL_570074\|2020-04 | ···················· | ···················· | ···················· | ···················· | ···················· | ···················· |

K 6902 hCoV-19/USA/WA-UW-5747/2020|EPI_ISL_570075|2020-04 ···················· ···················· ····················· ···················· ···················· ····················
K 6903 hCoV-19/USA/CT-UW-5775/2020|EPI_ISL_570076|2020-04 ···················· ···················· ····················· ···················· ···················· ····················
K 6904 hCoV-19/USA/WA-UW-5782/2020|EPI_ISL_570078|2020-04 ···················· ···················· ····················· ···················· ···················· ····················
K 6905 hCoV-19/USA/ID-UW-5888/2020|EPI_ISL_570079|2020-04 ···················· ···················· ····················· ···················· ···················· ····················
K 6906 hCoV-19/USA/WA-UW-589/2020|EPI_ISL_570080|2020-03- ···················· ···················· ····················· ···················· ···················· ····················
K 6907 hCoV-19/USA/WA-UW-603/2020|EPI_ISL_570081|2020-03- ···················· ···················· ····················· ···················· ···················· ····················
K 6908 hCoV-19/USA/WA-UW-6179/2020|EPI_ISL_570082|2020-04 ···················· ···················· ····················· ···················· ···················· ····················
K 6909 hCoV-19/USA/WA-UW-6181/2020|EPI_ISL_570083|2020-04 ···················· ···················· ····················· ···················· ···················· ····················
K 6910 hCoV-19/USA/WA-UW-6184/2020|EPI_ISL_570084|2020-04 ···················· ···················· ····················· ···················· ···················· ····················
K 6911 hCoV-19/USA/WA-UW-6190/2020|EPI_ISL_570086|2020-04 ···················· ···················· ····················· ···················· ···················· ····················
K 6912 hCoV-19/USA/WA-UW-6204/2020|EPI_ISL_570087|2020-04 ···················· ···················· ····················· ···················· ···················· ····················
K 6913 hCoV-19/Australia/VIC6716/2020|EPI_ISL_564007|2020 ···················· ···················· ····················· ···················· ···················· ····················
K 6914 hCoV-19/Australia/VIC6717/2020|EPI_ISL_564008|2020 ···················· ···················· ····················· ···················· ···················· ····················
K 6915 hCoV-19/Australia/VIC6719/2020|EPI_ISL_564010|2020 ···················· ····A··············· ····················· ···················· ···················· ····················
K 6916 hCoV-19/Australia/VIC6720/2020|EPI_ISL_564011|2020 ···················· ···················· ····················· ···················· ···················· ····················
K 6917 hCoV-19/Australia/VIC6721/2020|EPI_ISL_564012|2020 ···················· ···················· ····················· ···················· ···················· ····················
K 6918 hCoV-19/Australia/VIC6722/2020|EPI_ISL_564013|2020 ···················· ···················· ····················· ···················· ···················· ····················
K 6919 hCoV-19/Australia/VIC6725/2020|EPI_ISL_564014|2020 ···················· ···················· ····················· ···················· ···················· ····················
K 6920 hCoV-19/Australia/VIC6726/2020|EPI_ISL_564015|2020 ···················· ···················· ····················· ···················· ···················· ····················
K 6921 hCoV-19/Australia/VIC6748/2020|EPI_ISL_564027|2020 ···················· ···················· ····················· ···················· ···················· ····················
K 6922 hCoV-19/Australia/VIC6749/2020|EPI_ISL_564028|2020 ···················· ···················· ····················· ···················· ···················· ····················
K 6923 hCoV-19/Australia/VIC6751/2020|EPI_ISL_564029|2020 ···················· ···················· ····················· ···················· ···················· ····················
K 6924 hCoV-19/Australia/VIC6753/2020|EPI_ISL_564030|2020 ···················· ···················· ····················· ···················· ···················· ····················
K 6925 hCoV-19/Australia/VIC6755/2020|EPI_ISL_564031|2020 ···················· ···················· ····················· ···················· ···················· ····················
K 6926 hCoV-19/Australia/VIC6756/2020|EPI_ISL_564032|2020 ···················· ···················· ····················· ···················· ···················· ····················
K 6927 hCoV-19/Australia/VIC6757/2020|EPI_ISL_564033|2020 ···················· ···················· ····················· ···················· ···················· ····················
K 6928 hCoV-19/Australia/VIC6758/2020|EPI_ISL_564034|2020 ···················· ···················· ····················· ···················· ···················· ····················
K 6929 hCoV-19/Australia/VIC6759/2020|EPI_ISL_564035|2020 ···················· ···················· ····················· ···················· ···················· ····················
K 6930 hCoV-19/Australia/VIC6760/2020|EPI_ISL_564036|2020 ···················· ···················· ····················· ···················· ···················· ····················
K 6933 hCoV-19/Australia/VIC6763/2020|EPI_ISL_564039|2020 ···················· ···················· ····················· ···················· ···················· ····················
K 6934 hCoV-19/Australia/VIC6764/2020|EPI_ISL_564040|2020 ···················· ···················· ····················· ···················· ···················· ····················
K 6935 hCoV-19/Australia/VIC6765/2020|EPI_ISL_564041|2020 ···················· ···················· ····················· ···················· ···················· ····················
K 6936 hCoV-19/Australia/VIC6767/2020|EPI_ISL_564042|2020 ···················· ···················· ····················· ···················· ···················· ····················
K 6937 hCoV-19/Australia/VIC6768/2020|EPI_ISL_564043|2020 ···················· ···················· ····················· ···················· ···················· ····················
K 6938 hCoV-19/Australia/VIC6770/2020|EPI_ISL_564044|2020 ···················· ···················· ····················· ···················· ···················· ····················
K 6939 hCoV-19/Australia/VIC6772/2020|EPI_ISL_564045|2020 ···················· ···················· ····················· ···················· ···················· ····················
K 6940 hCoV-19/Australia/VIC6773/2020|EPI_ISL_564046|2020 ···················· ···················· ····················· ···················· ···················· ····················
K 6942 hCoV-19/Australia/VIC6775/2020|EPI_ISL_564048|2020 ···················· ···················· ····················· ···················· ···················· ····················
K 6943 hCoV-19/Australia/VIC6776/2020|EPI_ISL_564049|2020 ···················· ···················· ····················· ···················· ···················· ····················
K 6946 hCoV-19/Australia/VIC6782/2020|EPI_ISL_564052|2020 ···················· ···················· ····················· ···················· ···················· ····················
K 6947 hCoV-19/Australia/VIC6784/2020|EPI_ISL_564053|2020 ···················· ···················· ····················· ···················· ···················· ····················
K 6948 hCoV-19/Australia/VIC6786/2020|EPI_ISL_564054|2020 ···················· ···················· ····················· ···················· ···················· ····················
K 6949 hCoV-19/Australia/VIC6788/2020|EPI_ISL_564055|2020 ···················· ···················· ····················· ···················· ···················· ····················
K 6950 hCoV-19/Australia/VIC6789/2020|EPI_ISL_564056|2020 ···················· ···················· ···K················· ···················· ···················· ····················
K 6951 hCoV-19/Australia/VIC6790/2020|EPI_ISL_564057|2020 ···················· ···················· ····················· ···················· ···················· ····················
K 6952 hCoV-19/Australia/VIC6792/2020|EPI_ISL_564058|2020 ···················· ···················· ····················· ···················· ···················· ····················
K 6953 hCoV-19/Australia/VIC6793/2020|EPI_ISL_564059|2020 ···················· ···················· ····················· ···················· ···················· ····················
K 6954 hCoV-19/Australia/VIC6794/2020|EPI_ISL_564060|2020 ···················· ···················· ····················· ···················· ···················· ····················
K 6955 hCoV-19/Australia/VIC6795/2020|EPI_ISL_564061|2020 ···················· ···················· ····················· ···················· ···················· ····················
K 6956 hCoV-19/Australia/VIC6796/2020|EPI_ISL_564062|2020 ···················· ···················· ····················· ···················· ···················· ····················
K 6957 hCoV-19/Australia/VIC6797/2020|EPI_ISL_564063|2020 ···················· ···················· ····················· ···················· ···················· ····················
K 6958 hCoV-19/Australia/VIC6799/2020|EPI_ISL_564064|2020 ···················· ···················· ····················· ···················· ···················· ····················
K 6959 hCoV-19/Australia/VIC6800/2020|EPI_ISL_564065|2020 ···················· ···················· ····················· ···················· ···················· ····················
K 6960 hCoV-19/Australia/VIC6801/2020|EPI_ISL_564066|2020 ···················· ···················· ····················· ···················· ···················· ····················
K 6961 hCoV-19/Australia/VIC6804/2020|EPI_ISL_564067|2020 ···················· ···················· ····················· ···················· ···················· ····················
K 6962 hCoV-19/Australia/VIC6806/2020|EPI_ISL_564068|2020 ···················· ···················· ····················· ···················· ···················· ····················
K 6963 hCoV-19/Australia/VIC6807/2020|EPI_ISL_564069|2020 ···················· ···················· ····················· ···················· ···················· ····················
K 6964 hCoV-19/Australia/VIC6809/2020|EPI_ISL_564070|2020 ···················· ···················· ····················· ···················· ···················· ····················
K 6965 hCoV-19/Australia/VIC6812/2020|EPI_ISL_564071|2020 ···················· ···················· ····················· ···················· ···················· ····················
K 6966 hCoV-19/Australia/VIC6813/2020|EPI_ISL_564072|2020 ···················· ···················· ···K················· ···················· ···················· ····················
K 6967 hCoV-19/Australia/VIC6814/2020|EPI_ISL_564073|2020 ···················· ···················· ···K················· ···················· ···················· ····················
K 6968 hCoV-19/Australia/VIC6816/2020|EPI_ISL_564074|2020 ···················· ···················· ····················· ···················· ···················· ····················
K 6969 hCoV-19/Australia/VIC6817/2020|EPI_ISL_564075|2020 ···················· ···················· ····················· ···················· ···················· ····················
K 6972 hCoV-19/Australia/VIC6825/2020|EPI_ISL_564078|2020 ···················· ···················· ····················· ···················· ···················· ····················
K 6973 hCoV-19/Australia/VIC6826/2020|EPI_ISL_564079|2020 ···················· ···················· ····················· ···················· ···················· ····················
K 6974 hCoV-19/Australia/VIC6827/2020|EPI_ISL_564080|2020 ···················· ···················· ····················· ···················· ···················· ····················
K 6975 hCoV-19/Australia/VIC6829/2020|EPI_ISL_564081|2020 ···················· ···················· ····················· ···················· ···················· ····················
K 6976 hCoV-19/Australia/VIC6830/2020|EPI_ISL_564082|2020 ···················· ···················· ····················· ···················· ···················· ····················
K 6977 hCoV-19/Australia/VIC6831/2020|EPI_ISL_564083|2020 ···················· ···················· ····················· ···················· ···················· ····················
K 6978 hCoV-19/Australia/VIC6832/2020|EPI_ISL_564084|2020 ···················· ···················· ····················· ···················· ···················· ····················
K 6979 hCoV-19/Australia/VIC6833/2020|EPI_ISL_564085|2020 ···················· ···················· ····················· ···················· ···················· ····················
K 6981 hCoV-19/Australia/VIC6835/2020|EPI_ISL_564087|2020 ···················· ···················· ····················· ···················· ···················· ····················
K 6982 hCoV-19/Australia/VIC6836/2020|EPI_ISL_564088|2020 ···················· ···················· ····················· ···················· ···················· ····················
K 6983 hCoV-19/Australia/VIC6838/2020|EPI_ISL_564089|2020 ···················· ···················· ····················· ···················· ···················· ····················
K 6984 hCoV-19/Australia/VIC6857/2020|EPI_ISL_564100|2020 ···················· ···················· ····················· ···················· ···················· ····················
K 6985 hCoV-19/Australia/VIC6859/2020|EPI_ISL_564101|2020 ···················· ···················· ····················· ···················· ···················· ····················
K 6986 hCoV-19/Australia/VIC6862/2020|EPI_ISL_564102|2020 ···················· ···················· ····················· ···················· ···················· ····················
K 6987 hCoV-19/Australia/VIC6865/2020|EPI_ISL_564103|2020 ···················· ···················· ····················· ···················· ···················· ····················
K 6988 hCoV-19/Australia/VIC6883/2020|EPI_ISL_564115|2020 ···················· ···················· ····················· ···················· ···················· ····················
K 6989 hCoV-19/Australia/VIC6886/2020|EPI_ISL_564116|2020 ···················· ···················· ····················· ···················· ···················· ····················

| | | | | | | | | |
|---|---|---|---|---|---|---|---|---|
| K | 6990 | hCoV-19/Australia/VIC6890/2020\|EPI_ISL_564117\|2020 | ···················· | ···················· | ····················· | ···················· | ···················· | ···················· |
| K | 6991 | hCoV-19/Australia/VIC6891/2020\|EPI_ISL_564118\|2020 | ···················· | ···················· | ····················· | ···················· | ···················· | ···················· |
| K | 6992 | hCoV-19/Australia/VIC6893/2020\|EPI_ISL_564119\|2020 | ···················· | ···················· | ····················· | ···················· | ···················· | ···················· |
| K | 6993 | hCoV-19/Australia/VIC6897/2020\|EPI_ISL_564122\|2020 | ···················· | ···················· | ····················· | ···················· | ···················· | ···················· |
| K | 6994 | hCoV-19/Australia/VIC6898/2020\|EPI_ISL_564123\|2020 | ···················· | ···················· | ····················· | ···················· | ···················· | ···················· |
| K | 6995 | hCoV-19/Australia/VIC8988/2020\|EPI_ISL_565338\|2020 | ···················· | ···················· | ····················· | ···················· | ···················· | ···················· |
| K | 6996 | hCoV-19/Australia/VIC8989/2020\|EPI_ISL_565339\|2020 | ···················· | ···················· | ····················· | ···················· | ···················· | ···················· |
| K | 6997 | hCoV-19/Australia/VIC8990/2020\|EPI_ISL_565340\|2020 | ···················· | ···················· | ····················· | ···················· | ···················· | ···················· |
| K | 6998 | hCoV-19/Australia/VIC8991/2020\|EPI_ISL_565341\|2020 | ···················· | ···················· | ····················· | ···················· | ···················· | ···················· |
| K | 7000 | hCoV-19/Australia/VIC8993/2020\|EPI_ISL_565343\|2020 | ···················· | ·····A·············· | ····················· | ···················· | ···················· | ···················· |
| K | 7001 | hCoV-19/Australia/VIC8994/2020\|EPI_ISL_565344\|2020 | ···················· | ···················· | ····················· | ···················· | ···················· | ···················· |
| K | 7002 | hCoV-19/Australia/VIC8995/2020\|EPI_ISL_565345\|2020 | ···················· | ···················· | ····················· | ···················· | ···················· | ···················· |
| K | 7003 | hCoV-19/Australia/VIC8996/2020\|EPI_ISL_565346\|2020 | ···················· | ···················· | ····················· | ···················· | ···················· | ···················· |
| K | 7004 | hCoV-19/Australia/VIC9021/2020\|EPI_ISL_565358\|2020 | ···················· | ···················· | ····················· | ···················· | ···················· | ···················· |
| K | 7006 | hCoV-19/Australia/VIC9024/2020\|EPI_ISL_565360\|2020 | ···················· | ···················· | ····················· | ···················· | ···················· | ···················· |
| K | 7007 | hCoV-19/Australia/VIC9025/2020\|EPI_ISL_565361\|2020 | ···················· | ···················· | ····················· | ···················· | ···················· | ···················· |
| K | 7008 | hCoV-19/Australia/VIC9026/2020\|EPI_ISL_565362\|2020 | ···················· | ···················· | ····················· | ···················· | ···················· | ···················· |
| K | 7009 | hCoV-19/Australia/VIC9028/2020\|EPI_ISL_565363\|2020 | ···················· | ···················· | ····················· | ···················· | ···················· | ···················· |
| K | 7010 | hCoV-19/Australia/VIC9032/2020\|EPI_ISL_565364\|2020 | ···················· | ···················· | ····················· | ···················· | ···················· | ···················· |
| K | 7011 | hCoV-19/Australia/VIC9035/2020\|EPI_ISL_565365\|2020 | ···················· | ···················· | ····················· | ···················· | ···················· | ···················· |
| K | 7012 | hCoV-19/Australia/VIC9036/2020\|EPI_ISL_565366\|2020 | ···················· | ···················· | ····················· | ···················· | ···················· | ···················· |
| K | 7013 | hCoV-19/Australia/VIC9037/2020\|EPI_ISL_565367\|2020 | ···················· | ···················· | ····················· | ···················· | ···················· | ···················· |
| K | 7014 | hCoV-19/Australia/VIC9039/2020\|EPI_ISL_565368\|2020 | ···················· | ···················· | ····················· | ···················· | ···················· | ···················· |
| K | 7015 | hCoV-19/Australia/VIC9040/2020\|EPI_ISL_565369\|2020 | ···················· | ···················· | ····················· | ···················· | ···················· | ···················· |
| K | 7016 | hCoV-19/Australia/VIC9041/2020\|EPI_ISL_565370\|2020 | ···················· | ···················· | ····················· | ···················· | ···················· | ···················· |
| K | 7017 | hCoV-19/Australia/VIC9044/2020\|EPI_ISL_565371\|2020 | ···················· | ···················· | ····················· | ···················· | ···················· | ···················· |
| K | 7018 | hCoV-19/Australia/VIC9045/2020\|EPI_ISL_565372\|2020 | ···················· | ···················· | ····················· | ···················· | ···················· | ···················· |
| K | 7019 | hCoV-19/Australia/VIC9048/2020\|EPI_ISL_565373\|2020 | ···················· | ···················· | ····················· | ···················· | ···················· | ···················· |
| K | 7020 | hCoV-19/Australia/VIC9049/2020\|EPI_ISL_565374\|2020 | ···················· | ···················· | ····················· | ···················· | ···················· | ···················· |
| K | 7021 | hCoV-19/Australia/VIC9051/2020\|EPI_ISL_565375\|2020 | ···················· | ···················· | ····················· | ···················· | ···················· | ···················· |
| K | 7022 | hCoV-19/Australia/VIC9055/2020\|EPI_ISL_565376\|2020 | ···················· | ···················· | ····················· | ···················· | ···················· | ···················· |
| K | 7023 | hCoV-19/Australia/VIC9057/2020\|EPI_ISL_565377\|2020 | ···················· | ···················· | ····················· | ···················· | ···················· | ···················· |
| K | 7024 | hCoV-19/Australia/VIC9062/2020\|EPI_ISL_565378\|2020 | ···················· | ···················· | ····················· | ···················· | ···················· | ···················· |
| K | 7025 | hCoV-19/Australia/VIC9069/2020\|EPI_ISL_565379\|2020 | ···················· | ···················· | ····················· | ···················· | ···················· | ···················· |
| K | 7026 | hCoV-19/Australia/VIC9070/2020\|EPI_ISL_565380\|2020 | ···················· | ···················· | ····················· | ···················· | ···················· | ···················· |
| K | 7027 | hCoV-19/Australia/VIC9071/2020\|EPI_ISL_565381\|2020 | ···················· | ···················· | ····················· | ···················· | ···················· | ···················· |
| K | 7028 | hCoV-19/Australia/VIC9075/2020\|EPI_ISL_565382\|2020 | ···················· | ···················· | ····················· | ···················· | ···················· | ···················· |
| K | 7029 | hCoV-19/Australia/VIC9077/2020\|EPI_ISL_565383\|2020 | ···················· | ···················· | ····················· | ···················· | ···················· | ···················· |
| K | 7030 | hCoV-19/Australia/VIC9078/2020\|EPI_ISL_565384\|2020 | ···················· | ···················· | ····················· | ···················· | ···················· | ···················· |
| K | 7031 | hCoV-19/Australia/VIC9083/2020\|EPI_ISL_565385\|2020 | ···················· | ···················· | ····················· | ···················· | ···················· | ···················· |
| K | 7032 | hCoV-19/Australia/VIC9085/2020\|EPI_ISL_565386\|2020 | ···················· | ···················· | ····················· | ···················· | ···················· | ···················· |
| K | 7033 | hCoV-19/Australia/VIC9086/2020\|EPI_ISL_565387\|2020 | ···················· | ···················· | ····················· | ···················· | ···················· | ···················· |
| K | 7034 | hCoV-19/Australia/VIC9088/2020\|EPI_ISL_565388\|2020 | ···················· | ···················· | ····················· | ···················· | ···················· | ···················· |
| K | 7035 | hCoV-19/Australia/VIC9090/2020\|EPI_ISL_565389\|2020 | ···················· | ···················· | ····················· | ···················· | ···················· | ···················· |
| K | 7036 | hCoV-19/Australia/VIC9091/2020\|EPI_ISL_565390\|2020 | ···················· | ···················· | ····················· | ···················· | ···················· | ···················· |
| K | 7037 | hCoV-19/Australia/VIC9092/2020\|EPI_ISL_565391\|2020 | ···················· | ···················· | ····················· | ···················· | ···················· | ···················· |
| K | 7038 | hCoV-19/Australia/VIC9096/2020\|EPI_ISL_565392\|2020 | ···················· | ···················· | ····················· | ···················· | ···················· | ···················· |
| K | 7039 | hCoV-19/Australia/VIC9101/2020\|EPI_ISL_565393\|2020 | ···················· | ···················· | ····················· | ···················· | ···················· | ···················· |
| K | 7040 | hCoV-19/Australia/VIC9109/2020\|EPI_ISL_565394\|2020 | ···················· | ···················· | ····················· | ···················· | ···················· | ···················· |
| K | 7041 | hCoV-19/Australia/VIC9115/2020\|EPI_ISL_565395\|2020 | ···················· | ···················· | ····················· | ···················· | ···················· | ···················· |
| K | 7042 | hCoV-19/Australia/VIC9117/2020\|EPI_ISL_565396\|2020 | ···················· | ···················· | ···K················· | ···················· | ···················· | ···················· |
| K | 7043 | hCoV-19/Australia/VIC9119/2020\|EPI_ISL_565397\|2020 | ···················· | ···················· | ····················· | ···················· | ···················· | ···················· |
| K | 7044 | hCoV-19/Australia/VIC9122/2020\|EPI_ISL_565398\|2020 | ···················· | ···················· | ····················· | ···················· | ···················· | ···················· |
| K | 7045 | hCoV-19/Australia/VIC9125/2020\|EPI_ISL_565399\|2020 | ···················· | ···················· | ···K················· | ···················· | ···················· | ···················· |
| K | 7046 | hCoV-19/Australia/VIC9126/2020\|EPI_ISL_565400\|2020 | ···················· | ···················· | ····················· | ···················· | ···················· | ···················· |
| K | 7047 | hCoV-19/Australia/VIC9127/2020\|EPI_ISL_565401\|2020 | ···················· | ···················· | ···K················· | ···················· | ···················· | ···················· |
| K | 7048 | hCoV-19/Australia/VIC9131/2020\|EPI_ISL_565402\|2020 | ···················· | ···················· | ····················· | ···················· | ···················· | ···················· |
| K | 7049 | hCoV-19/Australia/VIC9133/2020\|EPI_ISL_565403\|2020 | ···················· | ···················· | ····················· | ···················· | ···················· | ···················· |
| K | 7050 | hCoV-19/Australia/VIC9134/2020\|EPI_ISL_565404\|2020 | ···················· | ···················· | ····················· | ···················· | ···················· | ···················· |
| K | 7051 | hCoV-19/Australia/VIC9135/2020\|EPI_ISL_565405\|2020 | ···················· | ···················· | ····················· | ···················· | ···················· | ···················· |
| K | 7052 | hCoV-19/Australia/VIC9136/2020\|EPI_ISL_565406\|2020 | ···················· | ···················· | ····················· | ···················· | ···················· | ···················· |
| K | 7053 | hCoV-19/Australia/VIC9140/2020\|EPI_ISL_565407\|2020 | ···················· | ···················· | ····················· | ···················· | ···················· | ···················· |
| K | 7054 | hCoV-19/Australia/VIC9141/2020\|EPI_ISL_565408\|2020 | ···················· | ···················· | ····················· | ···················· | ···················· | ···················· |
| K | 7055 | hCoV-19/Australia/VIC9142/2020\|EPI_ISL_565409\|2020 | ···················· | ···················· | ····················· | ···················· | ···················· | ···················· |
| K | 7056 | hCoV-19/Australia/VIC9143/2020\|EPI_ISL_565410\|2020 | ···················· | ···················· | ····················· | ···················· | ···················· | ···················· |
| K | 7057 | hCoV-19/Australia/VIC9144/2020\|EPI_ISL_565411\|2020 | ···················· | ···················· | ····················· | ···················· | ···················· | ···················· |
| K | 7058 | hCoV-19/Australia/VIC9145/2020\|EPI_ISL_565412\|2020 | ···················· | ···················· | ····················· | ···················· | ···················· | ···················· |
| K | 7059 | hCoV-19/Australia/VIC9146/2020\|EPI_ISL_565413\|2020 | ···················· | ···················· | ····················· | ···················· | ···················· | ···················· |
| K | 7060 | hCoV-19/Australia/VIC9147/2020\|EPI_ISL_565414\|2020 | ···················· | ···················· | ····················· | ···················· | ············R······· | ···················· |
| K | 7061 | hCoV-19/Australia/VIC9152/2020\|EPI_ISL_565415\|2020 | ···················· | ···················· | ····················· | ···················· | ···················· | ···················· |
| K | 7062 | hCoV-19/Australia/VIC9155/2020\|EPI_ISL_565416\|2020 | ···················· | ···················· | ····················· | ···················· | ···················· | ···················· |
| K | 7063 | hCoV-19/Australia/VIC9156/2020\|EPI_ISL_565417\|2020 | ···················· | ···················· | ····················· | ···················· | ···················· | ···················· |
| K | 7064 | hCoV-19/Australia/VIC9160/2020\|EPI_ISL_565418\|2020 | ···················· | ···················· | ····················· | ···················· | ···················· | ···················· |
| K | 7065 | hCoV-19/Australia/VIC9161/2020\|EPI_ISL_565419\|2020 | ···················· | ·····A·············· | ····················· | ···················· | ···················· | ···················· |
| K | 7066 | hCoV-19/Australia/VIC9163/2020\|EPI_ISL_565420\|2020 | ···················· | ···················· | ····················· | ···················· | ···················· | ···················· |
| K | 7067 | hCoV-19/Australia/VIC9167/2020\|EPI_ISL_565421\|2020 | ···················· | ···················· | ····················· | ···················· | ···················· | ···················· |
| K | 7068 | hCoV-19/Australia/VIC9171/2020\|EPI_ISL_565422\|2020 | ···················· | ···················· | ····················· | ···················· | ···················· | ···················· |
| K | 7069 | hCoV-19/Australia/VIC9175/2020\|EPI_ISL_565423\|2020 | ···················· | ···················· | ····················· | ···················· | ···················· | ···················· |
| K | 7070 | hCoV-19/Australia/VIC9177/2020\|EPI_ISL_565424\|2020 | ···················· | ···················· | ····················· | ···················· | ···················· | ···················· |
| K | 7071 | hCoV-19/Australia/VIC9178/2020\|EPI_ISL_565425\|2020 | ···················· | ···················· | ····················· | ···················· | ···················· | ···················· |

| | | | | | | | | |
|---|---|---|---|---|---|---|---|---|
| K | 7072 | hCoV-19/Australia/VIC9182/2020\|EPI_ISL_565426\|2020 | ···················· | ···················· | ···················· | ···················· | ···················· | ···················· |
| K | 7073 | hCoV-19/Australia/VIC9183/2020\|EPI_ISL_565427\|2020 | ···················· | ···················· | ···················· | ···················· | ···················· | ···················· |
| K | 7074 | hCoV-19/Australia/VIC9184/2020\|EPI_ISL_565428\|2020 | ···················· | ···················· | ···················· | ···················· | ···················· | ···················· |
| K | 7075 | hCoV-19/Australia/VIC9186/2020\|EPI_ISL_565429\|2020 | ···················· | ···················· | ···················· | ···················· | ···················· | ···················· |
| K | 7076 | hCoV-19/Australia/VIC9187/2020\|EPI_ISL_565430\|2020 | ···················· | ···················· | ···················· | ···················· | ···················· | ···················· |
| K | 7077 | hCoV-19/Australia/VIC9188/2020\|EPI_ISL_565431\|2020 | ···················· | ···················· | ···················· | ···················· | ···················· | ···················· |
| K | 7078 | hCoV-19/Australia/VIC9189/2020\|EPI_ISL_565432\|2020 | ···················· | ···················· | ···················· | ···················· | ···················· | ···················· |
| K | 7079 | hCoV-19/Australia/VIC9190/2020\|EPI_ISL_565433\|2020 | ···················· | ···················· | ···················· | ···················· | ···················· | ···················· |
| K | 7080 | hCoV-19/Australia/VIC9191/2020\|EPI_ISL_565434\|2020 | ···················· | ···················· | ···················· | ···················· | ···················· | ···················· |
| K | 7081 | hCoV-19/Australia/VIC9215/2020\|EPI_ISL_565446\|2020 | ···················· | ···················· | ···················· | ···················· | ···················· | ···················· |
| K | 7082 | hCoV-19/Australia/VIC9216/2020\|EPI_ISL_565447\|2020 | ···················· | ···················· | ···················· | ···················· | ···················· | ···················· |
| K | 7083 | hCoV-19/Australia/VIC9217/2020\|EPI_ISL_565448\|2020 | ···················· | ···················· | ···················· | ···················· | ···················· | ···················· |
| K | 7084 | hCoV-19/Australia/VIC9219/2020\|EPI_ISL_565449\|2020 | ···················· | ···················· | ···················· | ···················· | ···················· | ···················· |
| K | 7085 | hCoV-19/Australia/VIC9225/2020\|EPI_ISL_565453\|2020 | ···················· | ···················· | ···················· | ···················· | ···················· | ···················· |
| K | 7086 | hCoV-19/Australia/VIC9226/2020\|EPI_ISL_565454\|2020 | ···················· | ···················· | ···················· | ···················· | ···················· | ···················· |
| K | 7087 | hCoV-19/England/MILK-9C2E9E/2020\|EPI_ISL_566669\|20 | ···················· | ·A·················· | ···················· | ···················· | ···················· | ···A················ |
| K | 7088 | hCoV-19/England/MILK-9C2CFE/2020\|EPI_ISL_566670\|20 | ···················· | ···················· | ···················· | ···················· | ···················· | ···················· |
| K | 7089 | hCoV-19/England/MILK-9C1E80/2020\|EPI_ISL_566671\|20 | ···················· | ···················· | ···················· | ···················· | ···················· | ···················· |
| K | 7090 | hCoV-19/England/MILK-9C2371/2020\|EPI_ISL_566673\|20 | ···················· | ···················· | ···················· | ···················· | ···················· | ···················· |
| K | 7091 | hCoV-19/England/CAMC-9B1A8F/2020\|EPI_ISL_566674\|20 | ···················· | ···················· | ···················· | ···················· | ···················· | ···················· |
| K | 7092 | hCoV-19/England/MILK-9C26E1/2020\|EPI_ISL_566675\|20 | ···················· | ···················· | ···················· | ···················· | ···················· | ···················· |
| K | 7093 | hCoV-19/England/MILK-9C254E/2020\|EPI_ISL_566676\|20 | ···················· | ···················· | ···················· | ···················· | ······T············· | ···················· |
| K | 7094 | hCoV-19/England/MILK-9C27EE/2020\|EPI_ISL_566677\|20 | ···················· | ···················· | ···················· | ···················· | ···················· | ···················· |
| K | 7095 | hCoV-19/Wales/MILK-9C3808/2020\|EPI_ISL_566689\|2020 | ···················· | ···················· | ···················· | ···················· | ···················· | ···················· |
| K | 7096 | hCoV-19/England/MILK-9C32EC/2020\|EPI_ISL_566690\|20 | ···················· | ···················· | ···················· | ···················· | ···················· | ···················· |
| K | 7097 | hCoV-19/Wales/MILK-9C37ED/2020\|EPI_ISL_566691\|2020 | ···················· | ···················· | ···················· | ···················· | ···················· | ···················· |
| K | 7098 | hCoV-19/Wales/MILK-9C44B8/2020\|EPI_ISL_566692\|2020 | ···················· | ···················· | ···················· | ···················· | ···················· | ···················· |
| K | 7099 | hCoV-19/England/MILK-9C3C66/2020\|EPI_ISL_566693\|20 | ···················· | ···················· | ···················· | ···················· | ···················· | ···················· |
| K | 7100 | hCoV-19/England/MILK-9C35E3/2020\|EPI_ISL_566695\|20 | ···················· | ···················· | ···················· | ···················· | ···················· | ···················· |
| K | 7101 | hCoV-19/England/MILK-9C460D/2020\|EPI_ISL_566696\|20 | ···················· | ···················· | ···················· | ···················· | ···················· | ···················· |
| K | 7102 | hCoV-19/England/MILK-9C4500/2020\|EPI_ISL_566697\|20 | ···················· | ···················· | ···················· | ···················· | ···················· | ···················· |
| K | 7103 | hCoV-19/Wales/MILK-9C3DAF/2020\|EPI_ISL_566698\|2020 | ···················· | ···················· | ···················· | ···················· | ···················· | ···················· |
| K | 7104 | hCoV-19/England/MILK-9C453D/2020\|EPI_ISL_566699\|20 | ···················· | ···················· | ···················· | ···················· | ····T··············· | ···················· |
| K | 7105 | hCoV-19/England/MILK-9C3BF0/2020\|EPI_ISL_566700\|20 | ···················· | ·A·················· | ···················· | ···················· | ···················· | ···················· |
| K | 7106 | hCoV-19/England/MILK-9C3EF7/2020\|EPI_ISL_566701\|20 | ···················· | ···················· | ···················· | ···················· | ···················· | ···················· |
| K | 7107 | hCoV-19/England/MILK-9C3255/2020\|EPI_ISL_566702\|20 | ···················· | ······C············· | ···················· | ···················· | ···················· | ···················· |
| K | 7108 | hCoV-19/England/MILK-9C3835/2020\|EPI_ISL_566704\|20 | ···················· | ···················· | ···················· | ···················· | ···················· | ···················· |
| K | 7109 | hCoV-19/England/MILK-9C40E1/2020\|EPI_ISL_566706\|20 | ···················· | ···················· | ···················· | ···················· | ···················· | ···················· |
| K | 7110 | hCoV-19/England/MILK-9C3F9A/2020\|EPI_ISL_566708\|20 | ···················· | ···················· | ···················· | ···················· | ···················· | ···················· |
| K | 7111 | hCoV-19/England/MILK-9C36E0/2020\|EPI_ISL_566709\|20 | ···················· | ····A··············· | ···················· | ···················· | ···················· | ···················· |
| K | 7112 | hCoV-19/England/MILK-9C454C/2020\|EPI_ISL_566710\|20 | ···················· | ·A·················· | ···················· | ···················· | ···················· | ···················· |
| K | 7113 | hCoV-19/England/MILK-9C43CA/2020\|EPI_ISL_566711\|20 | ···················· | ···················· | ···················· | ···················· | ···················· | ···················· |
| K | 7114 | hCoV-19/Wales/MILK-9C3695/2020\|EPI_ISL_566712\|2020 | ···················· | ···················· | ···················· | ···················· | ···················· | ···················· |
| K | 7115 | hCoV-19/England/MILK-9C42FA/2020\|EPI_ISL_566713\|20 | ···················· | ·A·················· | ···················· | ···················· | ···················· | ···A················ |
| K | 7116 | hCoV-19/England/MILK-9C33F8/2020\|EPI_ISL_566714\|20 | ···················· | ···················· | ···················· | ···················· | ···················· | ···················· |
| K | 7117 | hCoV-19/England/MILK-9C34AA/2020\|EPI_ISL_566715\|20 | ···················· | ···················· | ···················· | ···················· | ···················· | ···················· |
| K | 7118 | hCoV-19/England/MILK-9C398D/2020\|EPI_ISL_566716\|20 | ···················· | ···················· | ···················· | ···················· | ···················· | ···················· |
| K | 7119 | hCoV-19/Wales/MILK-9C3932/2020\|EPI_ISL_566717\|2020 | ···················· | ···················· | ···················· | ···················· | ···················· | ···················· |
| K | 7120 | hCoV-19/England/MILK-9C41EE/2020\|EPI_ISL_566718\|20 | ···················· | ···················· | ···················· | ···················· | ···················· | ···················· |
| K | 7121 | hCoV-19/Wales/MILK-9C36C2/2020\|EPI_ISL_566719\|2020 | ···················· | ···················· | ···················· | ···················· | ···················· | ···················· |
| K | 7122 | hCoV-19/England/MILK-9C3738/2020\|EPI_ISL_566720\|20 | ···················· | ····A··············· | ···················· | ···················· | ···················· | ···················· |
| K | 7123 | hCoV-19/England/MILK-9C401E/2020\|EPI_ISL_566721\|20 | ···················· | ···················· | ···················· | ···················· | ···················· | ···················· |
| K | 7124 | hCoV-19/England/MILK-9C389F/2020\|EPI_ISL_566722\|20 | ···················· | ···················· | ·········T·········· | ···················· | ······T············· | ···················· |
| K | 7125 | hCoV-19/England/MILK-9C3194/2020\|EPI_ISL_566723\|20 | ···················· | ·A·················· | ···················· | ···················· | ···················· | ···A················ |
| K | 7126 | hCoV-19/England/MILK-9C462B/2020\|EPI_ISL_566725\|20 | ···················· | ···················· | ···················· | ···················· | ···················· | ···················· |
| K | 7127 | hCoV-19/England/MILK-9C4728/2020\|EPI_ISL_566726\|20 | ···················· | ···················· | ···················· | ···················· | ···················· | ···················· |
| K | 7128 | hCoV-19/England/MILK-9C3792/2020\|EPI_ISL_566727\|20 | ···················· | ·A·················· | ···················· | ···················· | ···················· | ···················· |
| K | 7129 | hCoV-19/England/MILK-9C451F/2020\|EPI_ISL_566728\|20 | ···················· | ·A·················· | ···················· | ···················· | ···················· | ···················· |
| K | 7130 | hCoV-19/Wales/MILK-9C3B78/2020\|EPI_ISL_566729\|2020 | ···················· | ···················· | ···················· | ···················· | ···················· | ···················· |
| K | 7131 | hCoV-19/Wales/MILK-9C3C57/2020\|EPI_ISL_566730\|2020 | ···················· | ···················· | ···················· | ···················· | ···················· | ···················· |
| K | 7132 | hCoV-19/England/MILK-9C39F6/2020\|EPI_ISL_566731\|20 | ···················· | ···················· | ···················· | ···················· | ···················· | ···················· |
| K | 7133 | hCoV-19/England/MILK-9C455B/2020\|EPI_ISL_566732\|20 | ···················· | ·A·················· | ···················· | ···················· | ···················· | ···A················ |
| K | 7134 | hCoV-19/England/MILK-9C31D0/2020\|EPI_ISL_566734\|20 | ···················· | ···················· | ···················· | ···················· | ···················· | ···················· |
| K | 7135 | hCoV-19/England/MILK-9C3B96/2020\|EPI_ISL_566735\|20 | ···················· | ···················· | ···················· | ···················· | ···················· | ···················· |
| K | 7136 | hCoV-19/England/MILK-9C3729/2020\|EPI_ISL_566736\|20 | ···················· | ···················· | ···················· | ···················· | ···················· | ···················· |
| K | 7137 | hCoV-19/England/MILK-9C3176/2020\|EPI_ISL_566737\|20 | ···················· | ···················· | ···················· | ···················· | ···················· | ···················· |
| K | 7138 | hCoV-19/England/MILK-9C33BC/2020\|EPI_ISL_566738\|20 | ···················· | ···················· | ···················· | ···················· | ···················· | ···················· |
| K | 7139 | hCoV-19/England/MILK-9C4227/2020\|EPI_ISL_566739\|20 | ···················· | ···················· | ···················· | ···················· | ···················· | ···················· |
| K | 7140 | hCoV-19/England/MILK-9C3756/2020\|EPI_ISL_566740\|20 | ···················· | ···················· | ···················· | ···················· | ···················· | ···················· |
| K | 7141 | hCoV-19/England/MILK-9C3E60/2020\|EPI_ISL_566741\|20 | ···················· | ···················· | ···················· | ···················· | ···················· | ···················· |
| K | 7142 | hCoV-19/England/MILK-9C30A6/2020\|EPI_ISL_566742\|20 | ···················· | ···················· | ···················· | ···················· | ···················· | ···················· |
| K | 7143 | hCoV-19/England/MILK-9C3361/2020\|EPI_ISL_566743\|20 | ···················· | ···················· | ···················· | ···················· | ···················· | ···················· |
| K | 7144 | hCoV-19/England/MILK-9C3B5A/2020\|EPI_ISL_566744\|20 | ···················· | ·A·················· | ···················· | ···················· | ···················· | ···················· |
| K | 7145 | hCoV-19/England/MILK-9C40A5/2020\|EPI_ISL_566745\|20 | ···················· | ···················· | ···················· | ···················· | ···················· | ···················· |
| K | 7146 | hCoV-19/England/MILK-9C461C/2020\|EPI_ISL_566748\|20 | ···················· | ···················· | ···················· | ···················· | ···················· | ···················· |
| K | 7147 | hCoV-19/England/MILK-9C3C39/2020\|EPI_ISL_566749\|20 | ···················· | ···················· | ···················· | ···················· | ···················· | ···················· |
| K | 7148 | hCoV-19/England/MILK-9C3880/2020\|EPI_ISL_566750\|20 | ···················· | ···················· | ···················· | ···················· | ···················· | ···················· |
| K | 7149 | hCoV-19/England/MILK-9C45E2/2020\|EPI_ISL_566751\|20 | ···················· | ···················· | ···················· | ···················· | ···················· | ···················· |
| K | 7150 | hCoV-19/England/MILK-9C449A/2020\|EPI_ISL_566752\|20 | ···················· | ···················· | ···················· | ···················· | ···················· | ···················· |
| K | 7151 | hCoV-19/England/MILK-9C3826/2020\|EPI_ISL_566753\|20 | ···················· | ···················· | ···················· | ···················· | ···················· | ···················· |

K 7152 hCoV-19/England/MILK-9C32A0/2020|EPI_ISL_566754|20 ···················· ···················· ····················· ···················· ···················· ····················
K 7153 hCoV-19/England/MILK-9C36D1/2020|EPI_ISL_566756|20 ···················· ····A··············· ····················· ···················· ···················· ····················
K 7154 hCoV-19/England/MILK-9C4209/2020|EPI_ISL_566757|20 ···················· ···················· ····················· ···················· ···················· ····················
K 7155 hCoV-19/England/MILK-9C38F9/2020|EPI_ISL_566758|20 ···················· ···················· ····················· ···················· ···················· ····················
K 7156 hCoV-19/England/MILK-9C3F30/2020|EPI_ISL_566759|20 ···················· ····A··············· ····················· ···················· ···················· ····················
K 7157 hCoV-19/England/MILK-9C40D2/2020|EPI_ISL_566760|20 ···················· ···················· ····················· ···················· ···················· ····················
K 7158 hCoV-19/England/MILK-9C4157/2020|EPI_ISL_566761|20 ···················· ·A·················· ····················· ···················· ···················· ···A················
K 7159 hCoV-19/England/MILK-9C38AE/2020|EPI_ISL_566762|20 ···················· ···················· ····················· ···················· ···················· ····················
K 7160 hCoV-19/England/MILK-9C3E42/2020|EPI_ISL_566763|20 ···················· ···················· ····················· ···················· ···················· ····················
K 7161 hCoV-19/England/MILK-9C42BE/2020|EPI_ISL_566764|20 ···················· ····A··············· ····················· ···················· ···················· ····················
K 7162 hCoV-19/England/MILK-9C3E8E/2020|EPI_ISL_566765|20 ···················· ···················· ····················· ···················· ···················· ····················
K 7163 hCoV-19/England/MILK-9C43D9/2020|EPI_ISL_566777|20 ···················· ···················· ····················· ···················· ···················· ····················
K 7164 hCoV-19/England/MILK-9C32DD/2020|EPI_ISL_566778|20 ···················· ···················· ····················· ···················· ······T············· ····················
K 7165 hCoV-19/England/MILK-9C34E6/2020|EPI_ISL_566779|20 ···················· ···················· ····················· ···················· ···················· ····················
K 7166 hCoV-19/England/MILK-9C3B69/2020|EPI_ISL_566784|20 ···················· ···················· ····················· ···················· ···················· ····················
K 7167 hCoV-19/England/MILK-9C34C8/2020|EPI_ISL_566785|20 ···················· ···················· ····················· ···················· ···················· ····················
K 7168 hCoV-19/England/ALDP-9BC79C/2020|EPI_ISL_567999|20 ···················· ···················· ····················· ···················· ···················· ····················
K 7169 hCoV-19/Lithuania/C20-05-R4/2020|EPI_ISL_560400|20 ···················· ···················· ····················· ···················· ···················· ····················
K 7170 hCoV-19/Lithuania/C20-05-R5/2020|EPI_ISL_560401|20 ···················· ···················· ····················· ···················· ···················· ····················
K 7171 hCoV-19/Lithuania/C20-05-R7/2020|EPI_ISL_560402|20 ···················· ···················· ····················· ···················· ···················· ····················
K 7172 hCoV-19/Lithuania/C20-05-R8/2020|EPI_ISL_560403|20 ···················· ···················· ····················· ···················· ···················· ····················
K 7173 hCoV-19/Lithuania/C20-05-R9/2020|EPI_ISL_560404|20 ···················· ···················· ····················· ···················· ···················· ····················
K 7174 hCoV-19/Lithuania/C20-06-R20/2020|EPI_ISL_560405|2 ···················· ···················· ····················· ···················· ···················· ····················
K 7175 hCoV-19/USA/DE-DHSS-F917779/2020|EPI_ISL_560406|20 ···················· ···················· ····················· ···················· ···················· ····················
K 7176 hCoV-19/Italy/CAM-IZSM-52479-129/2020|EPI_ISL_5604 ···················· ···················· ····················· ···················· ···················· ····················
K 7177 hCoV-19/Czech Republic/NRL-8121/2020|EPI_ISL_56040 ···················· ···················· ····················· ···················· ···················· ····················
K 7178 hCoV-19/Czech Republic/NRL-8122/2020|EPI_ISL_56040 ···················· ···················· ····················· ···················· ···················· ····················
K 7179 hCoV-19/Czech Republic/NRL-8123/2020|EPI_ISL_56041 ···················· ···················· ····················· ···················· ···················· ····················
K 7180 hCoV-19/Czech Republic/NRL-8124/2020|EPI_ISL_56041 ···················· ···················· ····················· ···················· ···················· ····················
K 7181 hCoV-19/Czech Republic/NRL-8125/2020|EPI_ISL_56041 ···················· ···················· ····················· ···················· ···················· ····················
K 7182 hCoV-19/Switzerland/AG-ETHZ-280072/2020|EPI_ISL_56 ···················· ···················· ····················· ···················· ······K············· ····················
K 7183 hCoV-19/Switzerland/AG-ETHZ-280079/2020|EPI_ISL_56 ···················· ···················· ····················· ···················· ······K············· ····················
K 7184 hCoV-19/Switzerland/AG-ETHZ-280083/2020|EPI_ISL_56 ···················· ···················· ····················· ···················· ···················· ····················
K 7185 hCoV-19/Switzerland/AG-ETHZ-280086/2020|EPI_ISL_56 ···················· ···················· ····················· ···················· ···················· ····················
K 7186 hCoV-19/Switzerland/AG-ETHZ-280113/2020|EPI_ISL_56 ···················· ···················· ····················· ···················· ···················· ···R················
K 7187 hCoV-19/Switzerland/AG-ETHZ-280134/2020|EPI_ISL_56 ···················· ···················· ····················· ···················· ···················· ····················
K 7188 hCoV-19/Switzerland/AG-ETHZ-280149/2020|EPI_ISL_56 ···················· ···················· ····················· ···················· ···················· ····················
K 7189 hCoV-19/Switzerland/AG-ETHZ-280156/2020|EPI_ISL_56 ···················· ···················· ····················· ···················· ······K············· ····················
K 7190 hCoV-19/Switzerland/AG-ETHZ-280178/2020|EPI_ISL_56 ···················· ···················· ····················· ···················· ···················· ····················
K 7191 hCoV-19/Switzerland/AG-ETHZ-280188/2020|EPI_ISL_56 ···················· ···················· ····················· ···················· ······K············· ····················
K 7192 hCoV-19/Switzerland/AG-ETHZ-280191/2020|EPI_ISL_56 ···················· ···················· ····················· ···················· ···················· ····················
K 7193 hCoV-19/Switzerland/AG-ETHZ-280247/2020|EPI_ISL_56 ···················· ···················· ····················· ···················· ···················· ····················
K 7194 hCoV-19/Switzerland/BE-ETHZ-280092/2020|EPI_ISL_56 ···················· ···················· ····················· ···················· ···················· ····················
K 7195 hCoV-19/Switzerland/BE-ETHZ-280094/2020|EPI_ISL_56 ···················· ···················· ····················· ···················· ···················· ····················
K 7196 hCoV-19/Switzerland/BE-ETHZ-280095/2020|EPI_ISL_56 ···················· ···················· ····················· ···················· ···················· ····················
K 7197 hCoV-19/Switzerland/BE-ETHZ-280109/2020|EPI_ISL_56 ···················· ···················· ····················· ···················· ···················· ····················
K 7198 hCoV-19/Switzerland/BE-ETHZ-280110/2020|EPI_ISL_56 ···················· ·A·················· ····················· ···················· ···················· ····················
K 7199 hCoV-19/Switzerland/BE-ETHZ-280116/2020|EPI_ISL_56 ···················· ·A·················· ····················· ···················· ···················· ····················
K 7200 hCoV-19/Switzerland/BE-ETHZ-280130/2020|EPI_ISL_56 ···················· ···················· ····················· ···················· ·······S············ ····················
K 7201 hCoV-19/Switzerland/BE-ETHZ-280132/2020|EPI_ISL_56 ···················· ···················· ····················· ···················· ···················· ····················
K 7202 hCoV-19/Switzerland/BE-ETHZ-280139/2020|EPI_ISL_56 ···················· ···················· ····················· ···················· ···················· ····················
K 7203 hCoV-19/Switzerland/BE-ETHZ-280140/2020|EPI_ISL_56 ···················· ···················· ····················· ···················· ···················· ····················
K 7204 hCoV-19/Switzerland/BE-ETHZ-280159/2020|EPI_ISL_56 ···················· ···················· ····················· ···················· ···················· ····················
K 7205 hCoV-19/USA/AK-PHL477/2020|EPI_ISL_560562|2020-06- ···················· ···················· ····················· ···················· ···················· ····················
K 7206 hCoV-19/USA/AK-PHL479/2020|EPI_ISL_560563|2020-06- ···················· ···················· ····················· ···················· ···················· ····················
K 7207 hCoV-19/USA/AK-PHL480/2020|EPI_ISL_560564|2020-06- ···················· ···················· ····················· ···················· ···················· ····················
K 7208 hCoV-19/USA/AK-PHL494/2020|EPI_ISL_560565|2020-06- ···················· ···················· ····················· ···················· ···················· ····················
K 7209 hCoV-19/USA/AK-PHL534/2020|EPI_ISL_560566|2020-06- ···················· ···················· ····················· ···················· ···················· ····················
K 7210 hCoV-19/France/CVL-8260/2020|EPI_ISL_560568|2020-0 ···················· ···················· ····················· ···················· ···················· ····················
K 7211 hCoV-19/France/CVL-8262/2020|EPI_ISL_560569|2020-0 ···················· ···················· ····················· ···················· ···················· ····················
K 7212 hCoV-19/France/CVL-8265/2020|EPI_ISL_560570|2020-0 ···················· ···················· ····················· ···················· ···················· ····················
K 7213 hCoV-19/France/CVL-8267/2020|EPI_ISL_560571|2020-0 ···················· ···················· ····················· ···················· ···················· ····················
K 7214 hCoV-19/France/CVL-8270/2020|EPI_ISL_560572|2020-0 ···················· ···················· ····················· ···················· ···················· ····················
K 7215 hCoV-19/France/CVL-8271/2020|EPI_ISL_560573|2020-0 ···················· ···················· ····················· ···················· ···················· ····················
K 7216 hCoV-19/France/CVL-8278/2020|EPI_ISL_560574|2020-0 ···················· ···················· ····················· ···················· ···················· ····················
K 7217 hCoV-19/France/CVL-8280/2020|EPI_ISL_560575|2020-0 ···················· ···················· ····················· ···················· ···················· ····················
K 7218 hCoV-19/France/CVL-8282/2020|EPI_ISL_560576|2020-0 ···················· ···················· ····················· ···················· ···················· ····················
K 7219 hCoV-19/France/CVL-8287/2020|EPI_ISL_560577|2020-0 ···················· ···················· ····················· ···················· ···················· ····················
K 7220 hCoV-19/France/CVL-8289/2020|EPI_ISL_560578|2020-0 ···················· ···················· ····················· ···················· ···················· ····················
K 7221 hCoV-19/France/CVL-8294/2020|EPI_ISL_560579|2020-0 ···················· ···················· ····················· ···················· ···················· ····················
K 7222 hCoV-19/France/CVL-8297/2020|EPI_ISL_560580|2020-0 ···················· ···················· ····················· ···················· ···················· ····················
K 7223 hCoV-19/France/GES-8500/2020|EPI_ISL_560581|2020-0 ···················· ···················· ····················· ···················· ···················· ····················
K 7224 hCoV-19/France/GES-8508/2020|EPI_ISL_560582|2020-0 ···················· ···················· ····················· ···················· ···················· ····················
K 7225 hCoV-19/France/GES-8509/2020|EPI_ISL_560583|2020-0 ···················· ···················· ····················· ···················· ···················· ····················
K 7226 hCoV-19/France/GES-8512/2020|EPI_ISL_560584|2020-0 ···················· ···················· ····················· ···················· ···················· ····················
K 7227 hCoV-19/France/GES-8517/2020|EPI_ISL_560585|2020-0 ···················· ···················· ····················· ···················· ···················· ····················
K 7228 hCoV-19/France/GES-8524/2020|EPI_ISL_560586|2020-0 ···················· ···················· ····················· ···················· ···················· ····················
K 7229 hCoV-19/France/GES-8536/2020|EPI_ISL_560587|2020-0 ···················· ···················· ····················· ···················· ···················· ····················
K 7230 hCoV-19/France/GES-8539/2020|EPI_ISL_560588|2020-0 ···················· ···················· ····················· ···················· ···················· ····················
K 7231 hCoV-19/France/GES-8541/2020|EPI_ISL_560589|2020-0 ···················· ···················· ····················· ···················· ···················· ····················

| | | | | | | | | | |
|---|---|---|---|---|---|---|---|---|---|
| K | 7232 | hCoV-19/France/GES-8544/2020\|EPI_ISL_560590\|2020-0 | ···················· | ···················· | ···················· | ···················· | ···················· | ···················· | |
| K | 7233 | hCoV-19/France/HDF-8231/2020\|EPI_ISL_560591\|2020-0 | ···················· | ···················· | ···················· | ···················· | ···················· | ···················· | |
| K | 7234 | hCoV-19/France/HDF-8233/2020\|EPI_ISL_560592\|2020-0 | ···················· | ···················· | ···················· | ···················· | ···················· | ···················· | |
| K | 7235 | hCoV-19/France/HDF-8236/2020\|EPI_ISL_560593\|2020-0 | ···················· | ···················· | ···················· | ···················· | ···················· | ···················· | |
| K | 7236 | hCoV-19/France/HDF-8242/2020\|EPI_ISL_560594\|2020-0 | ···················· | ···················· | ···················· | ···················· | ···················· | ···················· | |
| K | 7237 | hCoV-19/France/HDF-8244/2020\|EPI_ISL_560595\|2020-0 | ···················· | ···················· | ···················· | ···················· | ···················· | ···················· | |
| K | 7238 | hCoV-19/France/HDF-8249/2020\|EPI_ISL_560596\|2020-0 | ···················· | ···················· | ···················· | ···················· | ···················· | ···················· | |
| K | 7239 | hCoV-19/France/HDF-8255/2020\|EPI_ISL_560597\|2020-0 | ···················· | ···················· | ···················· | ···················· | ···················· | ···················· | |
| K | 7240 | hCoV-19/France/BRE-8930/2020\|EPI_ISL_560598\|2020-0 | ···················· | ···················· | ···················· | ···················· | ···················· | ···················· | |
| K | 7241 | hCoV-19/France/BRE-8931/2020\|EPI_ISL_560599\|2020-0 | ···················· | ···················· | ···················· | ···················· | ···················· | ···················· | |
| K | 7242 | hCoV-19/Australia/VIC6866/2020\|EPI_ISL_564104\|2020 | ···················· | ···················· | ···················· | ···················· | ···················· | ···················· | |
| K | 7243 | hCoV-19/Australia/VIC6867/2020\|EPI_ISL_564105\|2020 | ···················· | ···················· | ···················· | ···················· | ···················· | ···················· | |
| K | 7244 | hCoV-19/Australia/VIC6868/2020\|EPI_ISL_564106\|2020 | ···················· | ···················· | ···················· | ···················· | ···················· | ···················· | |
| K | 7245 | hCoV-19/Australia/VIC6869/2020\|EPI_ISL_564107\|2020 | ···················· | ···················· | ···················· | ···················· | ···················· | ···················· | |
| K | 7246 | hCoV-19/Australia/VIC6871/2020\|EPI_ISL_564108\|2020 | ···················· | ···················· | ···················· | ···················· | ···················· | ···················· | |
| K | 7247 | hCoV-19/Australia/VIC6875/2020\|EPI_ISL_564109\|2020 | ···················· | ···················· | ···················· | ···················· | ···················· | ···················· | |
| K | 7248 | hCoV-19/Australia/VIC6876/2020\|EPI_ISL_564110\|2020 | ···················· | ···················· | ···················· | ···················· | ···················· | ···················· | |
| K | 7249 | hCoV-19/Australia/VIC6878/2020\|EPI_ISL_564111\|2020 | ···················· | ···················· | ···················· | ···················· | ···················· | ···················· | |
| K | 7250 | hCoV-19/Australia/VIC6879/2020\|EPI_ISL_564112\|2020 | ···················· | ···················· | ···················· | ···················· | ···················· | ···················· | |
| K | 7251 | hCoV-19/Australia/VIC6881/2020\|EPI_ISL_564113\|2020 | ···················· | ···················· | ···················· | ···················· | ···················· | ···················· | |
| K | 7252 | hCoV-19/Australia/VIC6882/2020\|EPI_ISL_564114\|2020 | ···················· | ···················· | ···················· | ···················· | ···················· | ······NN············ | |
| K | 7253 | hCoV-19/Australia/VIC6894/2020\|EPI_ISL_564120\|2020 | ···················· | ···················· | ···················· | ···················· | ···················· | ···················· | |
| K | 7254 | hCoV-19/Australia/VIC6895/2020\|EPI_ISL_564121\|2020 | ···················· | ···················· | ···················· | ···················· | ···················· | ···················· | |
| K | 7255 | hCoV-19/Australia/VIC6899/2020\|EPI_ISL_564124\|2020 | ···················· | ···················· | ···················· | ···················· | ···················· | ···················· | |
| K | 7257 | hCoV-19/Australia/VIC8818/2020\|EPI_ISL_565215\|2020 | ···················· | ···················· | ···················· | ···················· | ···················· | ···················· | |
| K | 7258 | hCoV-19/Australia/VIC8819/2020\|EPI_ISL_565216\|2020 | ···················· | ···················· | ···················· | ···················· | ···················· | ···················· | |
| K | 7259 | hCoV-19/Australia/VIC8824/2020\|EPI_ISL_565217\|2020 | ···················· | ···················· | ···················· | ···················· | ···················· | ···················· | |
| K | 7260 | hCoV-19/Australia/VIC8825/2020\|EPI_ISL_565218\|2020 | ···················· | ···················· | ···················· | ···················· | ···················· | ···················· | |
| K | 7261 | hCoV-19/Australia/VIC8826/2020\|EPI_ISL_565219\|2020 | ···················· | ···················· | ···················· | ···················· | ···················· | ···················· | |
| K | 7262 | hCoV-19/Australia/VIC8828/2020\|EPI_ISL_565220\|2020 | ···················· | ···················· | ···················· | ···················· | ···················· | ···················· | |
| K | 7263 | hCoV-19/Australia/VIC8830/2020\|EPI_ISL_565221\|2020 | ···················· | ···················· | ···················· | ···················· | ···················· | ···················· | |
| K | 7264 | hCoV-19/Australia/VIC8831/2020\|EPI_ISL_565222\|2020 | ···················· | ···················· | ···················· | ···················· | ···················· | ···················· | |
| K | 7265 | hCoV-19/Australia/VIC8833/2020\|EPI_ISL_565223\|2020 | ···················· | ···················· | ···················· | ···················· | ···················· | ···················· | |
| K | 7266 | hCoV-19/Australia/VIC8834/2020\|EPI_ISL_565224\|2020 | ···················· | ···················· | ···················· | ···················· | ···················· | ···················· | |
| K | 7267 | hCoV-19/Australia/VIC8835/2020\|EPI_ISL_565225\|2020 | ···················· | ···················· | ···················· | ···················· | ···················· | ···················· | |
| K | 7268 | hCoV-19/Australia/VIC8837/2020\|EPI_ISL_565226\|2020 | ···················· | ·····A·············· | ···················· | ···················· | ···················· | ···················· | |
| K | 7269 | hCoV-19/Australia/VIC8840/2020\|EPI_ISL_565227\|2020 | ···················· | ···················· | ···················· | ···················· | ···················· | ···················· | |
| K | 7270 | hCoV-19/Australia/VIC8841/2020\|EPI_ISL_565228\|2020 | ···················· | ···················· | ···················· | ···················· | ···················· | ···················· | |
| K | 7271 | hCoV-19/Australia/VIC8842/2020\|EPI_ISL_565229\|2020 | ···················· | ···················· | ···················· | ···················· | ···················· | ···················· | |
| K | 7272 | hCoV-19/Australia/VIC8843/2020\|EPI_ISL_565230\|2020 | ···················· | ···················· | ···················· | ···················· | ···················· | ···················· | |
| K | 7273 | hCoV-19/Australia/VIC8845/2020\|EPI_ISL_565231\|2020 | ···················· | ···················· | ···················· | ···················· | ···················· | ···················· | |
| K | 7274 | hCoV-19/Australia/VIC8846/2020\|EPI_ISL_565232\|2020 | ···················· | ···················· | ···················· | ···················· | ···················· | ···················· | |
| K | 7275 | hCoV-19/Australia/VIC8847/2020\|EPI_ISL_565233\|2020 | ···················· | ···················· | ···················· | ···················· | ···················· | ···················· | |
| K | 7276 | hCoV-19/Australia/VIC8848/2020\|EPI_ISL_565234\|2020 | ···················· | ···················· | ···················· | ···················· | ···················· | ···················· | |
| K | 7277 | hCoV-19/Australia/VIC8850/2020\|EPI_ISL_565235\|2020 | ···················· | ···················· | ···················· | ···················· | ···················· | ···················· | |
| K | 7278 | hCoV-19/Australia/VIC8851/2020\|EPI_ISL_565236\|2020 | ···················· | ···················· | ···················· | ···················· | ···················· | ···················· | |
| K | 7279 | hCoV-19/Australia/VIC8852/2020\|EPI_ISL_565237\|2020 | ···················· | ···················· | ···················· | ···················· | ···················· | ···················· | |
| K | 7280 | hCoV-19/Australia/VIC8853/2020\|EPI_ISL_565238\|2020 | ···················· | ···················· | ···················· | ···················· | NNN················· | ···················· | 1 |
| K | 7281 | hCoV-19/Australia/VIC8854/2020\|EPI_ISL_565239\|2020 | ···················· | ···················· | ···················· | ···················· | ···················· | ···················· | |
| K | 7282 | hCoV-19/Australia/VIC8855/2020\|EPI_ISL_565240\|2020 | ···················· | ···················· | ···················· | ···················· | ···················· | ···················· | |
| K | 7283 | hCoV-19/Australia/VIC8856/2020\|EPI_ISL_565241\|2020 | ···················· | ···················· | ···················· | ···················· | ···················· | ···················· | |
| K | 7284 | hCoV-19/Australia/VIC8858/2020\|EPI_ISL_565242\|2020 | ···················· | ···················· | ···················· | ···················· | ···················· | ···················· | |
| K | 7285 | hCoV-19/Australia/VIC8860/2020\|EPI_ISL_565243\|2020 | ···················· | ···················· | ···················· | ···················· | ···················· | ···················· | |
| K | 7286 | hCoV-19/Australia/VIC8861/2020\|EPI_ISL_565244\|2020 | ···················· | ···················· | ···················· | ···················· | ···················· | ···················· | |
| K | 7287 | hCoV-19/Australia/VIC8864/2020\|EPI_ISL_565245\|2020 | ···················· | ···················· | ···················· | ···················· | ···················· | ···················· | |
| K | 7288 | hCoV-19/Australia/VIC8865/2020\|EPI_ISL_565246\|2020 | ···················· | ···················· | ···················· | ···················· | ···················· | ···················· | |
| K | 7289 | hCoV-19/Australia/VIC8866/2020\|EPI_ISL_565247\|2020 | ···················· | ···················· | ···················· | ···················· | ···················· | ···················· | |
| K | 7290 | hCoV-19/Australia/VIC8869/2020\|EPI_ISL_565248\|2020 | ···················· | ···················· | ···················· | ···················· | ···················· | ···················· | |
| K | 7291 | hCoV-19/Australia/VIC8870/2020\|EPI_ISL_565249\|2020 | ···················· | ···················· | ···················· | ···················· | ···················· | ···················· | |
| K | 7292 | hCoV-19/Australia/VIC8871/2020\|EPI_ISL_565250\|2020 | ···················· | ···················· | ···················· | ···················· | ···················· | ···················· | |
| K | 7293 | hCoV-19/Australia/VIC8872/2020\|EPI_ISL_565251\|2020 | ···················· | ···················· | ···················· | ···················· | ···················· | ···················· | |
| K | 7294 | hCoV-19/Australia/VIC8873/2020\|EPI_ISL_565252\|2020 | ···················· | ···················· | ···················· | ···················· | ···················· | ···················· | |
| K | 7295 | hCoV-19/Australia/VIC8874/2020\|EPI_ISL_565253\|2020 | ···················· | ···················· | ···················· | ···················· | ···················· | ···················· | |
| K | 7296 | hCoV-19/Australia/VIC8875/2020\|EPI_ISL_565254\|2020 | ···················· | ···················· | ···················· | ···················· | ···················· | ···················· | |
| K | 7297 | hCoV-19/Australia/VIC8876/2020\|EPI_ISL_565255\|2020 | ···················· | ···················· | ···················· | ···················· | ···················· | ···················· | |
| K | 7298 | hCoV-19/Australia/VIC8878/2020\|EPI_ISL_565256\|2020 | ···················· | ···················· | ···················· | ···················· | ···················· | ···················· | |
| K | 7299 | hCoV-19/Australia/VIC8879/2020\|EPI_ISL_565257\|2020 | ···················· | ···················· | ···················· | ···················· | ···················· | ···················· | |
| K | 7300 | hCoV-19/Australia/VIC8880/2020\|EPI_ISL_565258\|2020 | ···················· | ···················· | ···················· | ···················· | ···················· | ···················· | |
| K | 7301 | hCoV-19/Australia/VIC8883/2020\|EPI_ISL_565259\|2020 | ···················· | ·········A·········· | ···················· | ···················· | ···················· | ···················· | |
| K | 7302 | hCoV-19/Australia/VIC8884/2020\|EPI_ISL_565260\|2020 | ···················· | ···················· | ···················· | ···················· | ···················· | ···················· | |
| K | 7303 | hCoV-19/Australia/VIC8885/2020\|EPI_ISL_565261\|2020 | ···················· | ···················· | ···················· | ···················· | ···················· | ···················· | |
| K | 7304 | hCoV-19/Australia/VIC8886/2020\|EPI_ISL_565262\|2020 | ···················· | ···················· | ···················· | ···················· | ···················· | ···················· | |
| K | 7305 | hCoV-19/Australia/VIC8887/2020\|EPI_ISL_565263\|2020 | ···················· | ···················· | ···················· | ···················· | ···················· | ···················· | |
| K | 7306 | hCoV-19/Australia/VIC8889/2020\|EPI_ISL_565264\|2020 | ···················· | ···················· | ···················· | ···················· | ···················· | ···················· | |
| K | 7307 | hCoV-19/Australia/VIC8890/2020\|EPI_ISL_565265\|2020 | ···················· | ···················· | ···················· | ···················· | ···················· | ···················· | |
| K | 7308 | hCoV-19/Australia/VIC8892/2020\|EPI_ISL_565266\|2020 | ···················· | ···················· | ···················· | ···················· | ···················· | ···················· | |
| K | 7309 | hCoV-19/Australia/VIC8893/2020\|EPI_ISL_565267\|2020 | ···················· | ···················· | ···················· | ···················· | ···················· | ···················· | |
| K | 7310 | hCoV-19/Australia/VIC8894/2020\|EPI_ISL_565268\|2020 | ···················· | ···················· | ···················· | ···················· | ···················· | ···················· | |
| K | 7311 | hCoV-19/Australia/VIC8896/2020\|EPI_ISL_565269\|2020 | ···················· | ···················· | ···················· | ···················· | ···················· | ···················· | |
| K | 7312 | hCoV-19/Australia/VIC8897/2020\|EPI_ISL_565270\|2020 | ···················· | ···················· | ···················· | ···················· | ···················· | ···················· | |

| | | | | | | | | |
|---|---|---|---|---|---|---|---|---|
| K | 7313 | hCoV-19/Australia/VIC8899/2020\|EPI_ISL_565271\|2020 | ···················· | ···················· | ····················· | ···················· | ···················· | ···················· |
| K | 7314 | hCoV-19/Australia/VIC8900/2020\|EPI_ISL_565272\|2020 | ···················· | ···················· | ····················· | ···················· | ···················· | ···················· |
| K | 7315 | hCoV-19/Australia/VIC8902/2020\|EPI_ISL_565273\|2020 | ···················· | ···················· | ····················· | ···················· | ···················· | ···················· |
| K | 7316 | hCoV-19/Australia/VIC8903/2020\|EPI_ISL_565274\|2020 | ···················· | ···················· | ····················· | ···················· | ···················· | ···················· |
| K | 7317 | hCoV-19/Australia/VIC8904/2020\|EPI_ISL_565275\|2020 | ···················· | ···················· | ····················· | ···················· | ···················· | ···················· |
| K | 7318 | hCoV-19/Australia/VIC8905/2020\|EPI_ISL_565276\|2020 | ···················· | ···················· | ····················· | ···················· | ···················· | ···················· |
| K | 7319 | hCoV-19/Australia/VIC8908/2020\|EPI_ISL_565277\|2020 | ···················· | ···················· | ····················· | ···················· | ···················· | ···················· |
| K | 7320 | hCoV-19/Australia/VIC8910/2020\|EPI_ISL_565278\|2020 | ···················· | ···················· | ····················· | ···················· | ···················· | ···················· |
| K | 7321 | hCoV-19/Australia/VIC8911/2020\|EPI_ISL_565279\|2020 | ···················· | ···················· | ····················· | ···················· | ···················· | ···················· |
| K | 7322 | hCoV-19/Australia/VIC8945/2020\|EPI_ISL_565299\|2020 | ···················· | ···················· | ····················· | ···················· | ···················· | ···················· |
| K | 7323 | hCoV-19/Australia/VIC9193/2020\|EPI_ISL_565435\|2020 | ···················· | ···················· | ····················· | ···················· | ···················· | ···················· |
| K | 7324 | hCoV-19/Australia/VIC9194/2020\|EPI_ISL_565436\|2020 | ···················· | ···················· | ····················· | ···················· | ···················· | ···················· |
| K | 7325 | hCoV-19/Australia/VIC9195/2020\|EPI_ISL_565437\|2020 | ···················· | ···················· | ····················· | ···················· | ···················· | ···················· |
| K | 7326 | hCoV-19/Australia/VIC9196/2020\|EPI_ISL_565438\|2020 | ···················· | ···················· | ····················· | ···················· | ···················· | ···················· |
| K | 7327 | hCoV-19/Australia/VIC9201/2020\|EPI_ISL_565439\|2020 | ···················· | ···················· | ····················· | ···················· | ···················· | ···················· |
| K | 7328 | hCoV-19/Australia/VIC9202/2020\|EPI_ISL_565440\|2020 | ···················· | ···················· | ····················· | ···················· | ···················· | ···················· |
| K | 7329 | hCoV-19/Australia/VIC9204/2020\|EPI_ISL_565441\|2020 | ···················· | ···················· | ····················· | ···················· | ···················· | ···················· |
| K | 7330 | hCoV-19/Australia/VIC9208/2020\|EPI_ISL_565442\|2020 | ···················· | ···················· | ····················· | ···················· | ···················· | ···················· |
| K | 7331 | hCoV-19/Australia/VIC9211/2020\|EPI_ISL_565443\|2020 | ···················· | ···················· | ····················· | ···················· | ···················· | ···················· |
| K | 7332 | hCoV-19/Australia/VIC9212/2020\|EPI_ISL_565444\|2020 | ···················· | ···················· | ····················· | ···················· | ···················· | ···················· |
| K | 7333 | hCoV-19/Australia/VIC9213/2020\|EPI_ISL_565445\|2020 | ···················· | ···················· | ····················· | ···················· | ···················· | ···················· |
| K | 7334 | hCoV-19/Australia/VIC9220/2020\|EPI_ISL_565450\|2020 | ···················· | ···················· | ····················· | ···················· | ···················· | ···················· |
| K | 7335 | hCoV-19/Australia/VIC9222/2020\|EPI_ISL_565451\|2020 | ···················· | ···················· | ····················· | ···················· | ···················· | ···················· |
| K | 7336 | hCoV-19/Australia/VIC9223/2020\|EPI_ISL_565452\|2020 | ···················· | ···················· | ····················· | ···················· | ···················· | ···················· |
| K | 7337 | hCoV-19/Australia/VIC9231/2020\|EPI_ISL_565455\|2020 | ···················· | ···················· | ····················· | ···················· | ···················· | ···················· |
| K | 7338 | hCoV-19/Australia/VIC9232/2020\|EPI_ISL_565456\|2020 | ···················· | ···················· | ····················· | ···················· | ···················· | ···················· |
| K | 7339 | hCoV-19/England/MILK-9C27B1/2020\|EPI_ISL_566546\|20 | ···················· | ·A·················· | ····················· | ···················· | ···················· | ··A················· |
| K | 7340 | hCoV-19/England/MILK-9C2010/2020\|EPI_ISL_566547\|20 | ···················· | ·A·················· | ····················· | ···················· | ···················· | ···················· |
| K | 7341 | hCoV-19/England/MILK-9C220B/2020\|EPI_ISL_566548\|20 | ···················· | ·A·················· | ····················· | ···················· | ···················· | ···················· |
| K | 7342 | hCoV-19/Wales/MILK-9C2A03/2020\|EPI_ISL_566549\|2020 | ···················· | ···················· | ····················· | ···················· | ···················· | ···················· |
| K | 7343 | hCoV-19/England/MILK-9C260F/2020\|EPI_ISL_566550\|20 | ···················· | ···················· | ····················· | ···················· | ···················· | ···················· |
| K | 7344 | hCoV-19/England/MILK-9C2D73/2020\|EPI_ISL_566551\|20 | ···················· | ···················· | ····················· | ···················· | ···················· | ···················· |
| K | 7345 | hCoV-19/Wales/MILK-9C2256/2020\|EPI_ISL_566552\|2020 | ···················· | ···················· | ····················· | ···················· | ···················· | ···················· |
| K | 7346 | hCoV-19/Wales/MILK-9C26C3/2020\|EPI_ISL_566553\|2020 | ···················· | ···················· | ····················· | ···················· | ···················· | ···················· |
| K | 7347 | hCoV-19/England/MILK-9C2089/2020\|EPI_ISL_566555\|20 | ···················· | ···················· | ····················· | ···················· | ···················· | ···················· |
| K | 7348 | hCoV-19/England/MILK-9C2A5E/2020\|EPI_ISL_566556\|20 | ···················· | ···················· | ····················· | ···················· | ···················· | ···················· |
| K | 7349 | hCoV-19/England/MILK-9C2C2B/2020\|EPI_ISL_566557\|20 | ···················· | ···················· | ····················· | ···················· | ···················· | ···················· |
| K | 7350 | hCoV-19/England/MILK-9C1EF9/2020\|EPI_ISL_566558\|20 | ···················· | ···················· | ····················· | ···················· | ···················· | ···················· |
| K | 7351 | hCoV-19/England/MILK-9C23AE/2020\|EPI_ISL_566559\|20 | ···················· | ···················· | ····················· | ···················· | ···················· | ···················· |
| K | 7352 | hCoV-19/Wales/MILK-9C2423/2020\|EPI_ISL_566560\|2020 | ···················· | ···················· | ····················· | ···················· | ···················· | ···················· |
| K | 7353 | hCoV-19/England/MILK-9C2669/2020\|EPI_ISL_566561\|20 | ···················· | ···················· | ····················· | ···················· | ···················· | ···················· |
| K | 7354 | hCoV-19/England/MILK-9C2A7C/2020\|EPI_ISL_566562\|20 | ···················· | ·A·················· | ····················· | ···················· | ···················· | ··A················· |
| K | 7355 | hCoV-19/England/MILK-9C22B0/2020\|EPI_ISL_566564\|20 | ···················· | ···················· | ····················· | ···················· | ···················· | ···················· |
| K | 7356 | hCoV-19/England/MILK-9C2C94/2020\|EPI_ISL_566565\|20 | ···················· | ·A·················· | ····················· | ···················· | ···················· | ··A················· |
| K | 7357 | hCoV-19/Wales/MILK-9C2E43/2020\|EPI_ISL_566566\|2020 | ···················· | ···················· | ····················· | ···················· | ···················· | ···················· |
| K | 7358 | hCoV-19/England/CAMC-9B2417/2020\|EPI_ISL_566567\|20 | ···················· | ···················· | ····················· | ···················· | ···················· | ···················· |
| K | 7359 | hCoV-19/England/MILK-9C1FAB/2020\|EPI_ISL_566568\|20 | ···················· | ···················· | ····················· | ···················· | ···················· | ···················· |
| K | 7360 | hCoV-19/England/MILK-9C2757/2020\|EPI_ISL_566569\|20 | ···················· | ···················· | ····················· | ···················· | ···················· | ···················· |
| K | 7361 | hCoV-19/England/MILK-9C22DE/2020\|EPI_ISL_566570\|20 | ···················· | ···················· | ····················· | ···················· | ···················· | ···················· |
| K | 7362 | hCoV-19/Wales/MILK-9C20E3/2020\|EPI_ISL_566571\|2020 | ···················· | ···················· | ····················· | ···················· | ···················· | ···················· |
| K | 7363 | hCoV-19/Wales/MILK-9C1FE7/2020\|EPI_ISL_566572\|2020 | ···················· | ···················· | ····················· | ···················· | ···················· | ···················· |
| K | 7364 | hCoV-19/England/MILK-9C2344/2020\|EPI_ISL_566573\|20 | ···················· | ···················· | ····················· | ···················· | ···················· | ···················· |
| K | 7365 | hCoV-19/England/MILK-9C1F8D/2020\|EPI_ISL_566574\|20 | ···················· | ···················· | ····················· | ···················· | ···················· | ···················· |
| K | 7366 | hCoV-19/England/CAMC-9B1CC5/2020\|EPI_ISL_566576\|20 | ···················· | ···················· | ····················· | ···················· | ···················· | ···················· |
| K | 7367 | hCoV-19/England/MILK-9C2BD3/2020\|EPI_ISL_566577\|20 | ···················· | ···················· | ····················· | ···················· | ···················· | ···················· |
| K | 7368 | hCoV-19/England/MILK-9C2696/2020\|EPI_ISL_566578\|20 | ···················· | ···················· | ····················· | ···················· | ···················· | ···················· |
| K | 7369 | hCoV-19/England/MILK-9C1F32/2020\|EPI_ISL_566579\|20 | ···················· | ···················· | ····················· | ···················· | ···················· | ···················· |
| K | 7370 | hCoV-19/England/MILK-9C26F0/2020\|EPI_ISL_566580\|20 | ···················· | ···················· | ····················· | ···················· | ···················· | ···················· |
| K | 7371 | hCoV-19/England/MILK-9C2D46/2020\|EPI_ISL_566581\|20 | ···················· | ···················· | ····················· | ···················· | ···················· | ···················· |
| K | 7372 | hCoV-19/England/MILK-9C24E7/2020\|EPI_ISL_566582\|20 | ···················· | ···················· | ····················· | ···················· | ···················· | ···················· |
| K | 7373 | hCoV-19/England/MILK-9C27DF/2020\|EPI_ISL_566583\|20 | ···················· | ···················· | ····················· | ···················· | ···················· | ···················· |
| K | 7374 | hCoV-19/England/MILK-9C29D9/2020\|EPI_ISL_566584\|20 | ········T··········· | ···················· | ····················· | ···················· | ···················· | ···················· |
| K | 7375 | hCoV-19/Wales/MILK-9C2405/2020\|EPI_ISL_566585\|2020 | ···················· | ···················· | ····················· | ···················· | ···················· | ···················· |
| K | 7376 | hCoV-19/England/CAMC-9B1D0E/2020\|EPI_ISL_566586\|20 | ···················· | ···················· | ····················· | ···················· | ···················· | ···················· |
| K | 7377 | hCoV-19/England/CAMC-9B1B22/2020\|EPI_ISL_566589\|20 | ···················· | ···················· | ····················· | ···················· | ···················· | ···················· |
| K | 7378 | hCoV-19/England/MILK-9C2A4F/2020\|EPI_ISL_566590\|20 | ···················· | ······C············· | ····················· | ···················· | ···················· | ···················· |
| K | 7379 | hCoV-19/England/MILK-9C2432/2020\|EPI_ISL_566591\|20 | ···················· | ···················· | ····················· | ···················· | ···················· | ···················· |
| K | 7380 | hCoV-19/Wales/MILK-9C2F13/2020\|EPI_ISL_566592\|2020 | ···················· | ···················· | ····················· | ···················· | ···················· | ···················· |
| K | 7381 | hCoV-19/England/MILK-9C1FBA/2020\|EPI_ISL_566593\|20 | ···················· | ···················· | ····················· | ···················· | ···················· | ···················· |
| K | 7382 | hCoV-19/England/MILK-9C2A8B/2020\|EPI_ISL_566594\|20 | ···················· | ···················· | ····················· | ···················· | ···················· | ···················· |
| K | 7383 | hCoV-19/England/MILK-9C1E08/2020\|EPI_ISL_566595\|20 | ···················· | ···················· | ····················· | ···················· | ···················· | ···················· |
| K | 7384 | hCoV-19/England/MILK-9C1EBD/2020\|EPI_ISL_566596\|20 | ···················· | ···················· | ····················· | ···················· | ···················· | ···················· |
| K | 7385 | hCoV-19/England/MILK-9C2933/2020\|EPI_ISL_566597\|20 | ···················· | ···················· | ····················· | ···················· | ···················· | ···················· |
| K | 7386 | hCoV-19/England/MILK-9C2906/2020\|EPI_ISL_566598\|20 | ···················· | ···················· | ····················· | ···················· | ···················· | ···················· |
| K | 7387 | hCoV-19/England/MILK-9C2D64/2020\|EPI_ISL_566599\|20 | ···················· | ···················· | ····················· | ···················· | ···················· | ···················· |
| K | 7388 | hCoV-19/Wales/MILK-9C36A4/2020\|EPI_ISL_566766\|2020 | ···················· | ···················· | ····················· | ···················· | ···················· | ···················· |
| K | 7389 | hCoV-19/England/MILK-9C3273/2020\|EPI_ISL_566767\|20 | ···················· | ···················· | ····················· | ···················· | ···················· | ···················· |
| K | 7390 | hCoV-19/England/MILK-9C37A1/2020\|EPI_ISL_566768\|20 | ···················· | ···················· | ····················· | ···················· | ···················· | ···················· |
| K | 7391 | hCoV-19/England/MILK-9C35B6/2020\|EPI_ISL_566769\|20 | ···················· | ·A·······A·········· | ····················· | ···················· | ···················· | ···················· |
| K | 7392 | hCoV-19/England/MILK-9C370B/2020\|EPI_ISL_566770\|20 | ···················· | ···················· | ····················· | ···················· | ···················· | ···················· |

| | | | | | | | | |
|---|---|---|---|---|---|---|---|---|
| K | 7393 hCoV-19/Wales/MILK-9C3F12/2020\|EPI_ISL_566771\|2020 | ···················· | ···················· | ···················· | ···················· | ···················· | ···················· | |
| K | 7394 hCoV-19/England/MILK-9C3A99/2020\|EPI_ISL_566772\|20 | ···················· | ···················· | ···················· | ···················· | ···················· | ···················· | |
| K | 7395 hCoV-19/England/MILK-9C452E/2020\|EPI_ISL_566773\|20 | ···················· | ·A·················· | ···················· | ···················· | ···················· | ···················· | |
| K | 7396 hCoV-19/England/MILK-9C3BA5/2020\|EPI_ISL_566774\|20 | ···················· | ···················· | ···················· | ···················· | ···················· | ···················· | |
| K | 7397 hCoV-19/England/MILK-9C4351/2020\|EPI_ISL_566775\|20 | ···················· | ···················· | ···················· | ···················· | ···················· | ···················· | |
| K | 7398 hCoV-19/England/MILK-9C3F4F/2020\|EPI_ISL_566776\|20 | ···················· | ····A··············· | ···················· | ···················· | ···················· | ···················· | |
| K | 7399 hCoV-19/England/MILK-9C3EAC/2020\|EPI_ISL_566780\|20 | ···················· | ···················· | ···················· | ···················· | ···················· | ···················· | |
| K | 7400 hCoV-19/England/MILK-9C4579/2020\|EPI_ISL_566781\|20 | ···················· | ·A·················· | ···················· | ···················· | ···················· | ···A················ | |
| K | 7401 hCoV-19/England/MILK-9C3D90/2020\|EPI_ISL_566782\|20 | ···················· | ···················· | ···················· | ···················· | ···················· | ···················· | |
| K | 7402 hCoV-19/England/MILK-9C4148/2020\|EPI_ISL_566783\|20 | ···················· | ···················· | ···················· | ···················· | ···················· | ···················· | |
| K | 7403 hCoV-19/England/MILK-9C362C/2020\|EPI_ISL_566787\|20 | ···················· | ···················· | ···················· | ···················· | ···················· | ···················· | |
| K | 7404 hCoV-19/England/ALDP-9C50E0/2020\|EPI_ISL_567877\|20 | ···················· | ···················· | ···················· | ···················· | ···················· | ···················· | |
| K | 7405 hCoV-19/England/ALDP-9C5C37/2020\|EPI_ISL_567878\|20 | ···················· | ···················· | ···················· | ···················· | ···················· | ···················· | |
| K | 7406 hCoV-19/England/ALDP-9C5763/2020\|EPI_ISL_567879\|20 | ···················· | ···················· | ···················· | ···················· | ···················· | ···················· | |
| K | 7407 hCoV-19/England/ALDP-9C5EB9/2020\|EPI_ISL_567880\|20 | ···················· | ···················· | ···················· | ···················· | ···················· | ···················· | |
| K | 7408 hCoV-19/England/ALDP-9C5860/2020\|EPI_ISL_567881\|20 | ···················· | ···················· | ···················· | ···················· | ···················· | ···················· | |
| K | 7409 hCoV-19/England/ALDP-9C54E4/2020\|EPI_ISL_567882\|20 | ···················· | ···················· | ···················· | ···················· | ···················· | ···················· | |
| K | 7410 hCoV-19/England/ALDP-9C5FD4/2020\|EPI_ISL_567883\|20 | ···················· | ······A············· | ···················· | ···················· | ···················· | ···················· | |
| K | 7411 hCoV-19/England/ALDP-9C59D6/2020\|EPI_ISL_567884\|20 | ···················· | ···················· | ···················· | ···················· | ···················· | ···················· | |
| K | 7412 hCoV-19/England/ALDP-9C555A/2020\|EPI_ISL_567885\|20 | ···················· | ···················· | ···················· | ···················· | ···················· | ···················· | |
| K | 7413 hCoV-19/England/ALDP-9BE91C/2020\|EPI_ISL_567886\|20 | ···················· | ···················· | ···················· | ···················· | ···················· | ···················· | |
| K | 7414 hCoV-19/England/ALDP-9BDDE4/2020\|EPI_ISL_567888\|20 | ···················· | ···················· | ···················· | ···················· | ···················· | ···················· | |
| K | 7415 hCoV-19/England/ALDP-9BE062/2020\|EPI_ISL_567889\|20 | ···················· | ···················· | ···················· | ···················· | ···················· | ···················· | |
| K | 7416 hCoV-19/England/ALDP-9BE105/2020\|EPI_ISL_567890\|20 | ···················· | ····A··············· | ···················· | ···················· | ···················· | ···················· | |
| K | 7417 hCoV-19/Wales/ALDP-9BE84C/2020\|EPI_ISL_567891\|2020 | ···················· | ···················· | ···················· | ···················· | ···················· | ···················· | |
| K | 7418 hCoV-19/England/ALDP-9BDE0F/2020\|EPI_ISL_567892\|20 | ···················· | ···················· | ···················· | ···················· | ···················· | ···················· | |
| K | 7419 hCoV-19/England/ALDP-9BE554/2020\|EPI_ISL_567893\|20 | ···················· | ···················· | ···················· | ···················· | ···················· | ···················· | |
| K | 7420 hCoV-19/England/ALDP-9BDC32/2020\|EPI_ISL_567894\|20 | ···················· | ···················· | ···················· | ···················· | ···················· | ···················· | |
| K | 7421 hCoV-19/Wales/ALDP-9BE123/2020\|EPI_ISL_567895\|2020 | ···················· | ···················· | ···················· | ···················· | ···················· | ···················· | |
| K | 7422 hCoV-19/England/ALDP-9BC820/2020\|EPI_ISL_567896\|20 | ···················· | ···················· | ···················· | ···················· | ···················· | ···················· | |
| K | 7423 hCoV-19/England/ALDP-9BC6AE/2020\|EPI_ISL_567897\|20 | ···················· | ···················· | ···················· | ···················· | ···················· | ···················· | |
| K | 7424 hCoV-19/England/ALDP-9BE34B/2020\|EPI_ISL_567898\|20 | ···················· | ···················· | ···················· | ···················· | ···················· | ···················· | |
| K | 7425 hCoV-19/Switzerland/BE-ETHZ-280176/2020\|EPI_ISL_56 | ···················· | ···················· | ···················· | ···················· | ···················· | ···················· | |
| K | 7426 hCoV-19/Switzerland/BE-ETHZ-280185/2020\|EPI_ISL_56 | ···················· | ···················· | ···················· | ···················· | ···················· | ···················· | |
| K | 7427 hCoV-19/Switzerland/BE-ETHZ-280186/2020\|EPI_ISL_56 | ···················· | ···················· | ···················· | ···················· | ···················· | ···················· | |
| K | 7428 hCoV-19/Switzerland/BE-ETHZ-280193/2020\|EPI_ISL_56 | ···················· | ···················· | ···················· | ···················· | ···················· | ···················· | |
| K | 7429 hCoV-19/Switzerland/BE-ETHZ-280195/2020\|EPI_ISL_56 | ···················· | ···················· | ···················· | ···················· | ···················· | ···················· | |
| K | 7430 hCoV-19/Switzerland/BE-ETHZ-280208/2020\|EPI_ISL_56 | ···················· | ···················· | ···················· | ···················· | ···················· | ···················· | |
| K | 7431 hCoV-19/Switzerland/BE-ETHZ-280223/2020\|EPI_ISL_56 | ···················· | ···················· | ···················· | ···················· | ·······S············ | ···················· | |
| K | 7432 hCoV-19/Switzerland/BE-ETHZ-280224/2020\|EPI_ISL_56 | ···················· | ···················· | ···················· | ···················· | ···················· | ···················· | |
| K | 7433 hCoV-19/Switzerland/BE-ETHZ-280236/2020\|EPI_ISL_56 | ···················· | ···················· | ···················· | ···················· | ···················· | ···················· | |
| K | 7434 hCoV-19/Switzerland/BE-ETHZ-280239/2020\|EPI_ISL_56 | ···················· | ···················· | ···················· | ···················· | ···················· | ···················· | |
| K | 7435 hCoV-19/Switzerland/BE-ETHZ-280243/2020\|EPI_ISL_56 | ···················· | ···················· | ···················· | ···················· | ···················· | ···················· | |
| K | 7436 hCoV-19/Switzerland/BE-ETHZ-280245/2020\|EPI_ISL_56 | ···················· | ···················· | ···················· | ···················· | ···················· | ···················· | |
| K | 7437 hCoV-19/Switzerland/BL-ETHZ-280081/2020\|EPI_ISL_56 | ···················· | ···················· | ···················· | ···················· | ···················· | ···················· | |
| K | 7438 hCoV-19/Switzerland/BL-ETHZ-280082/2020\|EPI_ISL_56 | ···················· | ···················· | ···················· | ···················· | ···················· | ···················· | |
| K | 7439 hCoV-19/Switzerland/BL-ETHZ-280084/2020\|EPI_ISL_56 | ···················· | ···················· | ···················· | ···················· | ······K············· | ···················· | |
| K | 7440 hCoV-19/Switzerland/BL-ETHZ-280122/2020\|EPI_ISL_56 | ···················· | ···················· | ···················· | ···················· | ···················· | ·················Y·· | 1 |
| K | 7441 hCoV-19/Switzerland/BL-ETHZ-280150/2020\|EPI_ISL_56 | ···················· | ···················· | ···················· | ···················· | ···················· | ···················· | |
| K | 7442 hCoV-19/Switzerland/BL-ETHZ-280151/2020\|EPI_ISL_56 | ···················· | ···················· | ···················· | ···················· | ···················· | ···················· | |
| K | 7443 hCoV-19/Switzerland/BL-ETHZ-280152/2020\|EPI_ISL_56 | ···················· | ···················· | ···················· | ···················· | ···················· | ···················· | |
| K | 7444 hCoV-19/Switzerland/BL-ETHZ-280180/2020\|EPI_ISL_56 | ···················· | ···················· | ···················· | ···················· | ···················· | ···················· | |
| K | 7445 hCoV-19/Switzerland/BL-ETHZ-280184/2020\|EPI_ISL_56 | ···················· | ···················· | ···················· | ···················· | ···············Y···· | ···················· | |
| K | 7446 hCoV-19/Switzerland/BL-ETHZ-280187/2020\|EPI_ISL_56 | ···················· | ···················· | ···················· | ···················· | ···················· | ···················· | |
| K | 7447 hCoV-19/Switzerland/BL-ETHZ-280189/2020\|EPI_ISL_56 | ···················· | ···················· | ···················· | ···················· | ···················· | ···················· | |
| K | 7448 hCoV-19/Switzerland/BL-ETHZ-280190/2020\|EPI_ISL_56 | ···················· | ···················· | ···················· | ···················· | ···················· | ···················· | |
| K | 7449 hCoV-19/Switzerland/BL-ETHZ-280221/2020\|EPI_ISL_56 | ···················· | ···················· | ···················· | ···················· | ···················· | ···················· | |
| K | 7450 hCoV-19/Switzerland/BL-ETHZ-280232/2020\|EPI_ISL_56 | ···················· | ···················· | ···················· | ···················· | ···················· | ···················· | |
| K | 7451 hCoV-19/Switzerland/BS-ETHZ-280043/2020\|EPI_ISL_56 | ···················· | ·A·················· | ···················· | ···················· | ···················· | ···A················ | |
| K | 7452 hCoV-19/Switzerland/BS-ETHZ-280117/2020\|EPI_ISL_56 | ···················· | ···················· | ···················· | ···················· | ···················· | ···················· | |
| K | 7453 hCoV-19/Switzerland/BS-ETHZ-280226/2020\|EPI_ISL_56 | ···················· | ···················· | ···················· | ···················· | ···················· | ···················· | |
| K | 7454 hCoV-19/Switzerland/BS-ETHZ-280227/2020\|EPI_ISL_56 | ···················· | ···················· | ···················· | ···················· | ···················· | ···················· | |
| K | 7455 hCoV-19/Switzerland/GE-ETHZ-280201/2020\|EPI_ISL_56 | ···················· | ···················· | ···················· | ···················· | ···················· | ···················· | |
| K | 7456 hCoV-19/Switzerland/GR-ETHZ-280071/2020\|EPI_ISL_56 | ···················· | ···················· | ···················· | ···················· | ···················· | ···················· | |
| K | 7457 hCoV-19/Switzerland/GR-ETHZ-280120/2020\|EPI_ISL_56 | ···················· | ···················· | ···················· | ···················· | ···················· | ···················· | |
| K | 7458 hCoV-19/Switzerland/GR-ETHZ-280154/2020\|EPI_ISL_56 | ···················· | ···················· | ···················· | ···················· | ···················· | ···················· | |
| K | 7459 hCoV-19/Switzerland/SG-ETHZ-280233/2020\|EPI_ISL_56 | ···················· | ···················· | ···················· | ···················· | ···················· | ···················· | |
| K | 7460 hCoV-19/Switzerland/SZ-ETHZ-280121/2020\|EPI_ISL_56 | ···················· | ···················· | ···················· | ···················· | ···················· | ···················· | |
| K | 7461 hCoV-19/Switzerland/SZ-ETHZ-280242/2020\|EPI_ISL_56 | ···················· | ···················· | ···················· | ···················· | ···················· | ···················· | |
| K | 7462 hCoV-19/Switzerland/TI-ETHZ-280192/2020\|EPI_ISL_56 | ···················· | ···················· | ···················· | ···················· | ···················· | ···················· | |
| K | 7463 hCoV-19/Switzerland/TI-ETHZ-280194/2020\|EPI_ISL_56 | ···················· | ···················· | ···················· | ···················· | ···········M········ | ···················· | |
| K | 7464 hCoV-19/Switzerland/VD-ETHZ-280074/2020\|EPI_ISL_56 | ···················· | ···················· | ···················· | ···················· | ···················· | ···················· | |
| K | 7465 hCoV-19/Switzerland/VD-ETHZ-280075/2020\|EPI_ISL_56 | ···················· | ···················· | ···················· | ···················· | ···················· | ···················· | |
| K | 7466 hCoV-19/Switzerland/VD-ETHZ-280077/2020\|EPI_ISL_56 | ···················· | ···················· | ···················· | ···················· | ···················· | ···················· | |
| K | 7467 hCoV-19/Switzerland/VD-ETHZ-280099/2020\|EPI_ISL_56 | ···················· | ···················· | ···················· | ···················· | ···················· | ···················· | |
| K | 7468 hCoV-19/Switzerland/VD-ETHZ-280100/2020\|EPI_ISL_56 | ···················· | ···················· | ···················· | ···················· | ···················· | ···················· | |
| K | 7469 hCoV-19/Switzerland/VD-ETHZ-280101/2020\|EPI_ISL_56 | ···················· | ···················· | ···················· | ···················· | ···················· | ···················· | |
| K | 7470 hCoV-19/Switzerland/VD-ETHZ-280102/2020\|EPI_ISL_56 | ···················· | ···················· | ···················· | ···················· | ···················· | ···················· | |
| K | 7471 hCoV-19/Switzerland/VD-ETHZ-280103/2020\|EPI_ISL_56 | ···················· | ···················· | ···················· | ···················· | ···················· | ···················· | |
| K | 7472 hCoV-19/Switzerland/VD-ETHZ-280104/2020\|EPI_ISL_56 | ···················· | ···················· | ···················· | ···················· | ···················· | ···················· | |

| | | | | | | | | |
|---|---|---|---|---|---|---|---|---|
| K | 7473 hCoV-19/Switzerland/VD-ETHZ-280105/2020\|EPI_ISL_56 | ···················· | ···················· | ···················· | ···················· | ···················· | ···················· | |
| K | 7474 hCoV-19/Switzerland/VD-ETHZ-280106/2020\|EPI_ISL_56 | ···················· | ···················· | ···················· | ···················· | ···················· | ···················· | |
| K | 7475 hCoV-19/Switzerland/VD-ETHZ-280112/2020\|EPI_ISL_56 | ···················· | ···········R········ | ···················· | ···················· | ···············Y···· | ···················· | |
| K | 7476 hCoV-19/Switzerland/VD-ETHZ-280114/2020\|EPI_ISL_56 | ········Y··········· | ···················· | ·········K·········· | ···················· | ···················· | ···················· | |
| K | 7477 hCoV-19/Australia/VIC6698/2020\|EPI_ISL_564000\|2020 | ···················· | ···················· | ···················· | ···················· | ···················· | ···················· | |
| K | 7478 hCoV-19/Australia/VIC6701/2020\|EPI_ISL_564001\|2020 | ···················· | ···················· | ···················· | ···················· | ···················· | ···················· | |
| K | 7479 hCoV-19/Australia/VIC6706/2020\|EPI_ISL_564002\|2020 | ···················· | ···················· | ···K················ | ···················· | ···················· | ···················· | |
| K | 7480 hCoV-19/Australia/VIC6710/2020\|EPI_ISL_564003\|2020 | ···················· | ···················· | ···················· | ···················· | ···················· | ···················· | |
| K | 7481 hCoV-19/Australia/VIC6713/2020\|EPI_ISL_564004\|2020 | ···················· | ···················· | ···················· | ···················· | ···················· | ···················· | |
| K | 7482 hCoV-19/Australia/VIC8912/2020\|EPI_ISL_565280\|2020 | ···················· | ···················· | ···················· | ···················· | ···················· | ···················· | |
| K | 7483 hCoV-19/Australia/VIC8915/2020\|EPI_ISL_565281\|2020 | ···················· | ···················· | ···················· | ···················· | ···················· | ···················· | |
| K | 7484 hCoV-19/Australia/VIC8916/2020\|EPI_ISL_565282\|2020 | ···················· | ···················· | ···················· | ···················· | ···················· | ···················· | |
| K | 7485 hCoV-19/Australia/VIC8925/2020\|EPI_ISL_565283\|2020 | ···················· | ···················· | ···················· | ···················· | ···················· | ···················· | |
| K | 7487 hCoV-19/Australia/VIC8929/2020\|EPI_ISL_565285\|2020 | ···················· | ···················· | ···················· | ···················· | ···················· | ···················· | |
| K | 7488 hCoV-19/Australia/VIC8930/2020\|EPI_ISL_565286\|2020 | ···················· | ···················· | ···················· | ···················· | ···················· | ···················· | |
| K | 7489 hCoV-19/Australia/VIC8932/2020\|EPI_ISL_565287\|2020 | ···················· | ···················· | ···················· | ···················· | ···················· | ···················· | |
| K | 7490 hCoV-19/Australia/VIC8933/2020\|EPI_ISL_565288\|2020 | ···················· | ···················· | ···················· | ···················· | ···················· | ···················· | |
| K | 7491 hCoV-19/Australia/VIC8934/2020\|EPI_ISL_565289\|2020 | ···················· | ···················· | ···················· | ···················· | K··················· | ···················· | |
| K | 7492 hCoV-19/Australia/VIC8935/2020\|EPI_ISL_565290\|2020 | ···················· | ···················· | ···················· | ···················· | ···················· | ···················· | |
| K | 7493 hCoV-19/Australia/VIC8936/2020\|EPI_ISL_565291\|2020 | ···················· | ···················· | ···················· | ···················· | ···················· | ···················· | |
| K | 7494 hCoV-19/Australia/VIC8937/2020\|EPI_ISL_565292\|2020 | ···················· | ···················· | ···················· | ···················· | ···················· | ···················· | |
| K | 7495 hCoV-19/Australia/VIC8939/2020\|EPI_ISL_565293\|2020 | ···················· | ···················· | ···················· | ···················· | ···················· | ···················· | |
| K | 7496 hCoV-19/Australia/VIC8940/2020\|EPI_ISL_565294\|2020 | ···················· | ···················· | ···················· | ···················· | ···················· | ···················· | |
| K | 7497 hCoV-19/Australia/VIC8941/2020\|EPI_ISL_565295\|2020 | ···················· | ···················· | ···················· | ···················· | ···················· | ···················· | |
| K | 7498 hCoV-19/Australia/VIC8942/2020\|EPI_ISL_565296\|2020 | ···················· | ···················· | ···················· | ···················· | ···················· | ···················· | |
| K | 7499 hCoV-19/Australia/VIC8943/2020\|EPI_ISL_565297\|2020 | ···················· | ···················· | ···················· | ···················· | ··N················· | ···················· | |
| K | 7501 hCoV-19/Australia/VIC8946/2020\|EPI_ISL_565300\|2020 | ···················· | ···················· | ···················· | ···················· | ···················· | ···················· | |
| K | 7502 hCoV-19/Australia/VIC8947/2020\|EPI_ISL_565301\|2020 | ···················· | ···················· | ···················· | ···················· | ···················· | ···················· | |
| K | 7503 hCoV-19/Australia/VIC8948/2020\|EPI_ISL_565302\|2020 | ···················· | ···················· | ···················· | ···················· | ···················· | ···················· | |
| K | 7505 hCoV-19/Australia/VIC8950/2020\|EPI_ISL_565304\|2020 | ···················· | ···················· | ···················· | ···················· | ···················· | ···················· | |
| K | 7506 hCoV-19/Australia/VIC8951/2020\|EPI_ISL_565305\|2020 | ···················· | ···················· | ···················· | ···················· | ···················· | ···················· | |
| K | 7507 hCoV-19/Australia/VIC8952/2020\|EPI_ISL_565306\|2020 | ···················· | ···················· | ···················· | ···················· | ···················· | ···················· | |
| K | 7508 hCoV-19/Australia/VIC8953/2020\|EPI_ISL_565307\|2020 | ···················· | ···················· | ···················· | ···················· | ···················· | ···················· | |
| K | 7509 hCoV-19/Australia/VIC8954/2020\|EPI_ISL_565308\|2020 | ···················· | ···················· | ···················· | ···················· | ···················· | ···················· | |
| K | 7510 hCoV-19/Australia/VIC8955/2020\|EPI_ISL_565309\|2020 | ···················· | ···················· | ···················· | ···················· | ···················· | ···················· | |
| K | 7511 hCoV-19/Australia/VIC8957/2020\|EPI_ISL_565310\|2020 | ···················· | ···················· | ···················· | ···················· | ···················· | ···················· | |
| K | 7512 hCoV-19/Australia/VIC8959/2020\|EPI_ISL_565311\|2020 | ···················· | ···················· | ···················· | ···················· | ···················· | ···················· | |
| K | 7513 hCoV-19/Australia/VIC8960/2020\|EPI_ISL_565312\|2020 | ···················· | ···················· | ···················· | ···················· | ···················· | ···················· | |
| K | 7514 hCoV-19/Australia/VIC8961/2020\|EPI_ISL_565313\|2020 | ···················· | ···················· | ···················· | ···················· | ···················· | ···················· | |
| K | 7515 hCoV-19/Australia/VIC8962/2020\|EPI_ISL_565314\|2020 | ···················· | ···················· | ···················· | ···················· | ···················· | ···················· | |
| K | 7516 hCoV-19/Australia/VIC8963/2020\|EPI_ISL_565315\|2020 | ···················· | ···················· | ···················· | ···················· | ···················· | ···················· | |
| K | 7517 hCoV-19/Australia/VIC8964/2020\|EPI_ISL_565316\|2020 | ···················· | ···················· | ···················· | ···················· | ···················· | ···················· | |
| K | 7518 hCoV-19/Australia/VIC8965/2020\|EPI_ISL_565317\|2020 | ···················· | ···················· | ···················· | ···················· | ···················· | ···················· | |
| K | 7519 hCoV-19/Australia/VIC8966/2020\|EPI_ISL_565318\|2020 | ···················· | ···················· | ···················· | ···················· | ···················· | ···················· | |
| K | 7520 hCoV-19/Australia/VIC8967/2020\|EPI_ISL_565319\|2020 | ···················· | ···················· | ···················· | ···················· | ···················· | ···················· | |
| K | 7521 hCoV-19/Australia/VIC8968/2020\|EPI_ISL_565320\|2020 | ···················· | ···················· | ···················· | ···················· | ···················· | ···················· | |
| K | 7522 hCoV-19/Australia/VIC8969/2020\|EPI_ISL_565321\|2020 | ···················· | ···················· | ···················· | ···················· | ···················· | ···················· | |
| K | 7523 hCoV-19/Australia/VIC8970/2020\|EPI_ISL_565322\|2020 | ···················· | ···················· | ···················· | ···················· | ···················· | ···················· | |
| K | 7524 hCoV-19/Australia/VIC8971/2020\|EPI_ISL_565323\|2020 | ···················· | ···················· | ···················· | ···················· | ···················· | ···················· | |
| K | 7526 hCoV-19/Australia/VIC8974/2020\|EPI_ISL_565325\|2020 | ···················· | ···················· | ···················· | ···················· | ···················· | ···················· | |
| K | 7527 hCoV-19/Australia/VIC8975/2020\|EPI_ISL_565326\|2020 | ···················· | ···················· | ···················· | ···················· | NNN················· | ···················· | 1 |
| K | 7528 hCoV-19/Australia/VIC8977/2020\|EPI_ISL_565327\|2020 | ···················· | ···················· | ···················· | ···················· | ···················· | ···················· | |
| K | 7529 hCoV-19/Australia/VIC8978/2020\|EPI_ISL_565328\|2020 | ···················· | ···················· | ···················· | ···················· | ···················· | ···················· | |
| K | 7530 hCoV-19/Australia/VIC8979/2020\|EPI_ISL_565329\|2020 | ···················· | ···················· | ···················· | ···················· | ···················· | ···················· | |
| K | 7531 hCoV-19/Australia/VIC8980/2020\|EPI_ISL_565330\|2020 | ···················· | ···················· | ···················· | ···················· | ···················· | ···················· | |
| K | 7533 hCoV-19/Australia/VIC8982/2020\|EPI_ISL_565332\|2020 | ···················· | ···················· | ···················· | ···················· | ···················· | ···················· | |
| K | 7534 hCoV-19/Australia/VIC8983/2020\|EPI_ISL_565333\|2020 | ···················· | ···················· | ···················· | ···················· | ···················· | ···················· | |
| K | 7536 hCoV-19/Australia/VIC8985/2020\|EPI_ISL_565335\|2020 | ···················· | ···················· | ···················· | ···················· | NNN················· | ···················· | 1 |
| K | 7537 hCoV-19/England/MILK-9C2845/2020\|EPI_ISL_566600\|20 | ···················· | ···················· | ···················· | ···················· | ···················· | ···················· | |
| K | 7538 hCoV-19/England/MILK-9C2E52/2020\|EPI_ISL_566601\|20 | ···················· | ···················· | ···················· | ···················· | ···················· | ···················· | |
| K | 7539 hCoV-19/England/MILK-9C2C1C/2020\|EPI_ISL_566602\|20 | ···················· | ···················· | ···················· | ···················· | ···················· | ···················· | |
| K | 7540 hCoV-19/England/MILK-9C23DB/2020\|EPI_ISL_566603\|20 | ···················· | ···················· | ···················· | ···················· | ···················· | ···················· | |
| K | 7541 hCoV-19/England/MILK-9C26B4/2020\|EPI_ISL_566604\|20 | ···················· | ···················· | ···················· | ···················· | ···················· | ···················· | |
| K | 7542 hCoV-19/Wales/MILK-9C257B/2020\|EPI_ISL_566605\|2020 | ···················· | ···················· | ···················· | ···················· | ···················· | ···················· | |
| K | 7543 hCoV-19/England/MILK-9C2A21/2020\|EPI_ISL_566606\|20 | ···················· | ···················· | ···················· | ···················· | ···················· | ···················· | |
| K | 7544 hCoV-19/England/MILK-9C2B4C/2020\|EPI_ISL_566607\|20 | ···················· | ···················· | ···················· | ···················· | ······T············· | ···················· | |
| K | 7545 hCoV-19/Wales/MILK-9C256C/2020\|EPI_ISL_566608\|2020 | ···················· | ···················· | ···················· | ···················· | ···················· | ···················· | |
| K | 7546 hCoV-19/England/MILK-9C2380/2020\|EPI_ISL_566609\|20 | ···················· | ···················· | ···················· | ···················· | ···················· | ···················· | |
| K | 7547 hCoV-19/England/CAMC-9B1D95/2020\|EPI_ISL_566610\|20 | ···················· | ···················· | ···················· | ···················· | ···················· | ···················· | |
| K | 7548 hCoV-19/England/CAMC-9B1C3E/2020\|EPI_ISL_566611\|20 | ···················· | ···················· | ···················· | ···················· | ···················· | ···················· | |
| K | 7549 hCoV-19/England/MILK-9C21FF/2020\|EPI_ISL_566612\|20 | ···················· | ···················· | ···················· | ···················· | ···················· | ···················· | |
| K | 7550 hCoV-19/Wales/MILK-9C29F7/2020\|EPI_ISL_566613\|2020 | ···················· | ···················· | ···················· | ···················· | ···················· | ···················· | |
| K | 7551 hCoV-19/Wales/MILK-9C2FD7/2020\|EPI_ISL_566614\|2020 | ···················· | ···················· | ···················· | ···················· | ···················· | ···················· | |
| K | 7552 hCoV-19/England/MILK-9C2924/2020\|EPI_ISL_566615\|20 | ···················· | ···················· | ···················· | ···················· | ···················· | ···················· | |
| K | 7553 hCoV-19/England/MILK-9C29AC/2020\|EPI_ISL_566616\|20 | ···················· | ·A·················· | ···················· | ···················· | ···················· | ···A················ | |
| K | 7554 hCoV-19/England/MILK-9C213B/2020\|EPI_ISL_566617\|20 | ···················· | ···················· | ···················· | ···················· | ···················· | ···················· | |
| K | 7555 hCoV-19/England/MILK-9C2915/2020\|EPI_ISL_566618\|20 | ···················· | ···················· | ···················· | ···················· | ···················· | ···················· | |
| K | 7556 hCoV-19/England/MILK-9C2C49/2020\|EPI_ISL_566620\|20 | ···················· | ···················· | ···················· | ···················· | ···················· | ···················· | |
| K | 7557 hCoV-19/England/MILK-9C2D0A/2020\|EPI_ISL_566621\|20 | ···················· | ···················· | ···················· | ···················· | ···················· | ···················· | |
| K | 7558 hCoV-19/England/MILK-9C202F/2020\|EPI_ISL_566622\|20 | ···················· | ···················· | ···················· | ···················· | ···················· | ···················· | |

K 7559 hCoV-19/England/MILK-9C2E61/2020|EPI_ISL_566623|20 ···················· ···················· ····················· ···················· ···················· ····················
K 7560 hCoV-19/England/MILK-9C2B79/2020|EPI_ISL_566624|20 ···················· ···················· ····················· ···················· ···················· ····················
K 7561 hCoV-19/England/CAMC-9B2839/2020|EPI_ISL_566625|20 ···················· ···················· ····················· ···················· ···················· ····················
K 7562 hCoV-19/Wales/MILK-9C255D/2020|EPI_ISL_566626|2020 ···················· ···················· ····················· ···················· ···················· ····················
K 7563 hCoV-19/England/MILK-9C25D5/2020|EPI_ISL_566627|20 ···················· ···················· ····················· ···················· ···················· ····················
K 7564 hCoV-19/England/MILK-9C2317/2020|EPI_ISL_566628|20 ···················· ······C············· ····················· ···················· ···················· ····················
K 7565 hCoV-19/England/MILK-9C25C6/2020|EPI_ISL_566629|20 ···················· ···················· ····················· ···················· ···················· ····················
K 7566 hCoV-19/England/MILK-9C2D82/2020|EPI_ISL_566630|20 ···················· ···················· ····················· ···················· ···················· ····················
K 7567 hCoV-19/England/MILK-9C271B/2020|EPI_ISL_566632|20 ···················· ···················· ····················· ···················· ···················· ····················
K 7568 hCoV-19/England/MILK-9C2265/2020|EPI_ISL_566633|20 ···················· ···················· ····················· ···················· ···················· ····················
K 7569 hCoV-19/England/MILK-9C2809/2020|EPI_ISL_566634|20 ···················· ···················· ·········T··········· ···················· ······T············· ····················
K 7570 hCoV-19/Wales/MILK-9C2247/2020|EPI_ISL_566636|2020 ···················· ···················· ····················· ···················· ···················· ····················
K 7571 hCoV-19/England/CAMC-9B1B31/2020|EPI_ISL_566638|20 ···················· ···················· ····················· ···················· ···················· ····················
K 7572 hCoV-19/England/MILK-9C2836/2020|EPI_ISL_566639|20 ···················· ···················· ····················· ···················· ······T············· ····················
K 7573 hCoV-19/England/MILK-9C264B/2020|EPI_ISL_566640|20 ···················· ···················· ····················· ···················· ···················· ····················
K 7574 hCoV-19/England/MILK-9C26A5/2020|EPI_ISL_566641|20 ···················· ···················· ····················· ···················· ···················· ····················
K 7575 hCoV-19/England/MILK-9C2326/2020|EPI_ISL_566642|20 ···················· ···················· ····················· ···················· ···················· ····················
K 7576 hCoV-19/England/MILK-9C2DFB/2020|EPI_ISL_566643|20 ···················· ···················· ····················· ···················· ···················· ····················
K 7577 hCoV-19/England/MILK-9C21B3/2020|EPI_ISL_566644|20 ···················· ···················· ····················· ···················· ···················· ····················
K 7578 hCoV-19/Wales/MILK-9C2335/2020|EPI_ISL_566645|2020 ···················· ···················· ····················· ···················· ···················· ····················
K 7579 hCoV-19/England/MILK-9C2AB8/2020|EPI_ISL_566646|20 ···················· ···················· ····················· ···················· ···················· ····················
K 7580 hCoV-19/England/MILK-9C262D/2020|EPI_ISL_566649|20 ···················· ···················· ····················· ···················· ···················· ····················
K 7581 hCoV-19/Wales/MILK-9C1F50/2020|EPI_ISL_566650|2020 ···················· ···················· ····················· ···················· ···················· ····················
K 7582 hCoV-19/England/MILK-9C205C/2020|EPI_ISL_566651|20 ···················· ···················· ····················· ···················· ···················· ····················
K 7583 hCoV-19/England/MILK-9C2739/2020|EPI_ISL_566652|20 ···················· ···················· ····················· ···················· ···················· ····················
K 7584 hCoV-19/England/MILK-9C1E26/2020|EPI_ISL_566653|20 ···················· ···················· ····················· ···················· ···················· ····················
K 7585 hCoV-19/Wales/MILK-9C21D1/2020|EPI_ISL_566654|2020 ···················· ···················· ····················· ···················· ···················· ····················
K 7586 hCoV-19/England/MILK-9C214A/2020|EPI_ISL_566655|20 ···················· ···················· ····················· ···················· ···················· ····················
K 7587 hCoV-19/England/MILK-9C1FF6/2020|EPI_ISL_566656|20 ···················· ···················· ····················· ···················· ···················· ····················
K 7588 hCoV-19/England/MILK-9C25A8/2020|EPI_ISL_566657|20 ···················· ···················· ····················· ···················· ···················· ····················
K 7589 hCoV-19/England/MILK-9C253F/2020|EPI_ISL_566658|20 ···················· ·A·················· ····················· ···················· ···················· ···A················
K 7590 hCoV-19/England/CAMC-9B1C4D/2020|EPI_ISL_566659|20 ···················· ·A·················· ····················· ···················· ···················· ···A················
K 7591 hCoV-19/England/MILK-9C2BA6/2020|EPI_ISL_566660|20 ···················· ···················· ····················· ···················· ······T············· ····················
K 7592 hCoV-19/England/CAMC-9B1AF8/2020|EPI_ISL_566661|20 ···················· ···················· ····················· ···················· ···················· ····················
K 7593 hCoV-19/England/CAMC-9B22A4/2020|EPI_ISL_566662|20 ···················· ·A·················· ····················· ···················· ···················· ···A················
K 7594 hCoV-19/Wales/MILK-9C1DFC/2020|EPI_ISL_566663|2020 ···················· ···················· ····················· ···················· ···················· ····················
K 7595 hCoV-19/Wales/MILK-9C2EDA/2020|EPI_ISL_566664|2020 ···················· ···················· ····················· ···················· ···················· ····················
K 7596 hCoV-19/England/MILK-9C1F23/2020|EPI_ISL_566665|20 ···················· ·A·················· ····················· ···················· ···················· ···A················
K 7597 hCoV-19/England/CAMC-9B1C10/2020|EPI_ISL_566666|20 ···················· ···················· ····················· ···················· ···················· ····················
K 7598 hCoV-19/England/ALDP-9BDBEA/2020|EPI_ISL_567903|20 ···················· ···················· ····················· ···················· ···················· ····················
K 7599 hCoV-19/England/ALDP-9BDE4B/2020|EPI_ISL_567904|20 ···················· ···················· ····················· ···················· ···················· ····················
K 7600 hCoV-19/England/ALDP-9BEAA0/2020|EPI_ISL_567905|20 ···················· ···················· ····················· ···················· ···················· ····················
K 7601 hCoV-19/England/ALDP-9BC705/2020|EPI_ISL_567906|20 ···················· ···················· ····················· ···················· ···················· ····················
K 7602 hCoV-19/Wales/ALDP-9BE1D8/2020|EPI_ISL_567907|2020 ···················· ···················· ····················· ···················· ···················· ····················
K 7603 hCoV-19/England/ALDP-9BEACE/2020|EPI_ISL_567908|20 ···················· ···················· ····················· ···················· ···················· ····················
K 7604 hCoV-19/England/ALDP-9BE2A8/2020|EPI_ISL_567909|20 ···················· ···················· ····················· ···················· ···················· ····················
K 7605 hCoV-19/England/ALDP-9BE82E/2020|EPI_ISL_567910|20 ···················· ···················· ····················· ···················· ···················· ····················
K 7606 hCoV-19/England/ALDP-9BDFA2/2020|EPI_ISL_567912|20 ···················· ···················· ····················· ···················· ···················· ····················
K 7607 hCoV-19/England/ALDP-9BE75E/2020|EPI_ISL_567913|20 ···················· ···················· ····················· ···················· ···················· ····················
K 7608 hCoV-19/England/ALDP-9BC6EA/2020|EPI_ISL_567915|20 ···················· ···················· ····················· ···················· ···················· ····················
K 7609 hCoV-19/England/ALDP-9BE457/2020|EPI_ISL_567916|20 ···················· ···················· ····················· ···················· ···················· ····················
K 7610 hCoV-19/England/ALDP-9BE24E/2020|EPI_ISL_567918|20 ···················· ···················· ····················· ···················· ···················· ····················
K 7611 hCoV-19/England/ALDP-9BE4C0/2020|EPI_ISL_567919|20 ···················· ···················· ····················· ···················· ···················· ····················
K 7612 hCoV-19/England/ALDP-9BC7F6/2020|EPI_ISL_567920|20 ···················· ···················· ····················· ···················· ···················· ····················
K 7613 hCoV-19/England/ALDP-9BE994/2020|EPI_ISL_567921|20 ···················· ···················· ····················· ···················· ···················· ····················
K 7614 hCoV-19/England/ALDP-9BDA92/2020|EPI_ISL_567922|20 ···················· ···················· ····················· ···················· ···················· ····················
K 7615 hCoV-19/England/ALDP-9BDFDF/2020|EPI_ISL_567923|20 ···················· ···················· ····················· ···················· ···················· ····················
K 7616 hCoV-19/England/ALDP-9BE800/2020|EPI_ISL_567924|20 ···················· ···················· ····················· ···················· ···················· ····················
K 7617 hCoV-19/England/ALDP-9BE448/2020|EPI_ISL_567925|20 ···················· ···················· ····················· ···················· ···················· ····················
K 7618 hCoV-19/England/ALDP-9BEBAD/2020|EPI_ISL_567926|20 ···················· ·A·······A·········· ····················· ···················· ···················· ····················
K 7619 hCoV-19/England/ALDP-9BDCE7/2020|EPI_ISL_567927|20 ···················· ···················· ····················· ···················· ···················· ····················
K 7620 hCoV-19/England/ALDP-9BE484/2020|EPI_ISL_567928|20 ···················· ···················· ····················· ···················· ···················· ····················
K 7621 hCoV-19/England/ALDP-9BDACF/2020|EPI_ISL_567929|20 ···················· ···················· ····················· ···················· ···················· ····················
K 7622 hCoV-19/England/ALDP-9BE2E4/2020|EPI_ISL_567930|20 ···················· ···················· ····················· ···················· ···················· ····················
K 7623 hCoV-19/England/ALDP-9BDBDB/2020|EPI_ISL_567931|20 ···················· ···················· ····················· ···················· ···················· ····················
K 7624 hCoV-19/England/ALDP-9BE967/2020|EPI_ISL_567932|20 ···················· ···················· ····················· ···················· ···················· ····················
K 7625 hCoV-19/England/ALDP-9BE035/2020|EPI_ISL_567933|20 ···················· ···················· ····················· ···················· ···················· ····················
K 7626 hCoV-19/England/ALDP-9BE33C/2020|EPI_ISL_567935|20 ···················· ···················· ····················· ···················· ···················· ····················
K 7627 hCoV-19/England/ALDP-9BD9B3/2020|EPI_ISL_567936|20 ···················· ···················· ····················· ···················· ···················· ····················
K 7628 hCoV-19/England/ALDP-9BE19C/2020|EPI_ISL_567937|20 ···················· ···················· ····················· ···················· ···················· ····················
K 7629 hCoV-19/England/ALDP-9BE8F1/2020|EPI_ISL_567938|20 ···················· ···················· ····················· ···················· ···················· ····················
K 7630 hCoV-19/England/ALDP-9BD9FF/2020|EPI_ISL_567939|20 ···················· ···················· ····················· ···················· ···················· ····················
K 7631 hCoV-19/England/ALDP-9BDA65/2020|EPI_ISL_567940|20 ···················· ···················· ····················· ···················· ···················· ····················
K 7632 hCoV-19/England/ALDP-9BEB43/2020|EPI_ISL_567942|20 ···················· ···················· ····················· ···················· ···················· ····················
K 7633 hCoV-19/England/ALDP-9BDA1A/2020|EPI_ISL_567943|20 ···················· ···················· ····················· ···················· ···················· ····················
K 7634 hCoV-19/Wales/ALDP-9BE92B/2020|EPI_ISL_567944|2020 ···················· ···················· ····················· ···················· ···················· ····················
K 7635 hCoV-19/England/ALDP-9BC8C6/2020|EPI_ISL_567945|20 ···················· ···················· ····················· ···················· ···················· ····················
K 7636 hCoV-19/England/ALDP-9BDFFD/2020|EPI_ISL_567946|20 ···················· ···················· ····················· ···················· ···················· ····················
K 7637 hCoV-19/England/ALDP-9BDF1B/2020|EPI_ISL_567947|20 ···················· ·A·················· ····················· ···················· ···················· ····················
K 7638 hCoV-19/England/ALDP-9BDB71/2020|EPI_ISL_567948|20 ···················· ···················· ····················· ···················· ···················· ····················

| | | | | | | | | |
|---|---|---|---|---|---|---|---|---|
| K | 7639 | hCoV-19/England/ALDP-9BDBCC/2020\|EPI_ISL_567949\|20 | .................... | .................... | .................... | .................... | .................... | .................... |
| K | 7640 | hCoV-19/England/ALDP-9BC662/2020\|EPI_ISL_567950\|20 | .................... | .................... | .................... | .................... | .................... | .................... |
| K | 7641 | hCoV-19/England/ALDP-9BE18D/2020\|EPI_ISL_567951\|20 | .................... | .................... | .................... | .................... | .................... | .................... |
| K | 7642 | hCoV-19/England/ALDP-9BDF93/2020\|EPI_ISL_567952\|20 | .................... | .................... | .................... | .................... | .................... | .................... |
| K | 7643 | hCoV-19/England/ALDP-9BE949/2020\|EPI_ISL_567953\|20 | .................... | .A.................. | .................... | .................... | .................... | ..A................. |
| K | 7644 | hCoV-19/England/ALDP-9BDE78/2020\|EPI_ISL_567955\|20 | .................... | .................... | .................... | .................... | .................... | .................... |
| K | 7645 | hCoV-19/England/ALDP-9BE27B/2020\|EPI_ISL_567957\|20 | .................... | .................... | .................... | .................... | .................... | .................... |
| K | 7646 | hCoV-19/England/ALDP-9BE563/2020\|EPI_ISL_567959\|20 | .................... | .................... | .................... | .................... | .................... | .................... |
| K | 7647 | hCoV-19/England/ALDP-9BDC05/2020\|EPI_ISL_567960\|20 | .................... | .................... | .................... | .................... | .................... | .................... |
| K | 7648 | hCoV-19/England/ALDP-9BDA29/2020\|EPI_ISL_567961\|20 | .................... | .................... | .................... | .................... | .................... | .................... |
| K | 7649 | hCoV-19/England/ALDP-9BE439/2020\|EPI_ISL_567962\|20 | .................... | .A.................. | .................... | .................... | .................... | ..A................. |
| K | 7650 | hCoV-19/England/ALDP-9BE6D9/2020\|EPI_ISL_567963\|20 | .................... | .................... | .................... | .................... | .................... | .................... |
| K | 7651 | hCoV-19/England/ALDP-9BDD20/2020\|EPI_ISL_567964\|20 | .................... | .................... | .................... | .................... | .................... | .................... |
| K | 7652 | hCoV-19/England/ALDP-9BE79A/2020\|EPI_ISL_567965\|20 | .................... | .................... | .................... | .................... | .................... | .................... |
| K | 7653 | hCoV-19/England/ALDP-9BE4DF/2020\|EPI_ISL_567966\|20 | .................... | .................... | .................... | .................... | .................... | .................... |
| K | 7654 | hCoV-19/England/ALDP-9BE1E7/2020\|EPI_ISL_567967\|20 | .................... | .................... | .................... | .................... | .................... | .................... |
| K | 7655 | hCoV-19/England/ALDP-9BDD4E/2020\|EPI_ISL_567968\|20 | .................... | .................... | .................... | .................... | .................... | .................... |
| K | 7656 | hCoV-19/England/ALDP-9BE044/2020\|EPI_ISL_567969\|20 | .................... | .................... | .................... | .................... | .................... | .................... |
| K | 7657 | hCoV-19/England/ALDP-9BE3B4/2020\|EPI_ISL_567971\|20 | .................... | .................... | .................... | .................... | .................... | .................... |
| K | 7658 | hCoV-19/England/ALDP-9BE42A/2020\|EPI_ISL_567973\|20 | .................... | .................... | .................... | .................... | .................... | .................... |
| K | 7659 | hCoV-19/England/ALDP-9BDD99/2020\|EPI_ISL_567974\|20 | .................... | .................... | .................... | .................... | .................... | .................... |
| K | 7660 | hCoV-19/England/ALDP-9BDC9C/2020\|EPI_ISL_567975\|20 | .................... | .................... | .................... | .................... | .................... | .................... |
| K | 7661 | hCoV-19/England/ALDP-9BE30F/2020\|EPI_ISL_567976\|20 | .................... | .................... | .................... | .................... | .................... | .................... |
| K | 7662 | hCoV-19/England/ALDP-9BDC14/2020\|EPI_ISL_567977\|20 | .................... | .................... | .................... | .................... | .................... | .................... |
| K | 7663 | hCoV-19/England/ALDP-9BEDE3/2020\|EPI_ISL_567978\|20 | .................... | .................... | .................... | .................... | .................... | .................... |
| K | 7664 | hCoV-19/England/ALDP-9BDB53/2020\|EPI_ISL_567979\|20 | .................... | .................... | .................... | .................... | .................... | .................... |
| K | 7665 | hCoV-19/England/ALDP-9BDE3C/2020\|EPI_ISL_567980\|20 | .................... | .................... | .................... | .................... | .................... | .................... |
| K | 7666 | hCoV-19/England/ALDP-9BE017/2020\|EPI_ISL_567981\|20 | .................... | .................... | .................... | .................... | .................... | .................... |
| K | 7667 | hCoV-19/England/ALDP-9BDB80/2020\|EPI_ISL_567982\|20 | .................... | .................... | .................... | .................... | .................... | .................... |
| K | 7668 | hCoV-19/England/ALDP-9BEDA7/2020\|EPI_ISL_567983\|20 | .................... | .................... | .................... | .................... | .................... | .................... |
| K | 7669 | hCoV-19/England/ALDP-9BED98/2020\|EPI_ISL_567985\|20 | .................... | .................... | .................... | .................... | .................... | .................... |
| K | 7670 | hCoV-19/England/ALDP-9BEAFB/2020\|EPI_ISL_567986\|20 | .................... | .................... | .................... | .................... | .................... | .................... |
| K | 7671 | hCoV-19/England/ALDP-9BC94B/2020\|EPI_ISL_567987\|20 | .................... | .................... | .................... | .................... | .................... | .................... |
| K | 7672 | hCoV-19/England/ALDP-9BEB25/2020\|EPI_ISL_567988\|20 | .................... | .................... | .................... | .................... | .................... | .................... |
| K | 7673 | hCoV-19/England/ALDP-9BC6BD/2020\|EPI_ISL_567989\|20 | .................... | .................... | .................... | .................... | .................... | .................... |
| K | 7674 | hCoV-19/England/ALDP-9BED3E/2020\|EPI_ISL_567990\|20 | .................... | .................... | .................... | .................... | .................... | .................... |
| K | 7675 | hCoV-19/England/ALDP-9BE3F0/2020\|EPI_ISL_567992\|20 | .................... | .................... | .................... | .................... | .................... | .................... |
| K | 7676 | hCoV-19/England/ALDP-9BE78B/2020\|EPI_ISL_567993\|20 | .................... | .................... | .................... | .................... | .................... | .................... |
| K | 7677 | hCoV-19/England/ALDP-9BEC6E/2020\|EPI_ISL_567994\|20 | .................... | .A.................. | .................... | .................... | .................... | ..A................. |
| K | 7678 | hCoV-19/England/ALDP-9BDAFC/2020\|EPI_ISL_567995\|20 | .................... | .................... | .................... | .................... | .................... | .................... |
| K | 7679 | hCoV-19/England/ALDP-9BE888/2020\|EPI_ISL_567996\|20 | .................... | .................... | .................... | .................... | .................... | .................... |
| K | 7680 | hCoV-19/England/ALDP-9BE008/2020\|EPI_ISL_567997\|20 | .................... | .................... | .................... | .................... | .................... | .................... |
| K | 7681 | hCoV-19/Australia/VIC10398/2020\|EPI_ISL_562900\|202 | .................... | .................... | .................... | .................... | .................... | .................... |
| K | 7682 | hCoV-19/Australia/VIC10401/2020\|EPI_ISL_562901\|202 | .................... | .................... | .................... | .................... | .................... | .................... |
| K | 7683 | hCoV-19/Australia/VIC10402/2020\|EPI_ISL_562902\|202 | .................... | .................... | .................... | .................... | .................... | .................... |
| K | 7684 | hCoV-19/Australia/VIC10403/2020\|EPI_ISL_562903\|202 | .................... | .................... | .................... | .................... | .................... | .................... |
| K | 7685 | hCoV-19/Australia/VIC10406/2020\|EPI_ISL_562904\|202 | .................... | .................... | .................... | .................... | .................... | .................... |
| K | 7691 | hCoV-19/Australia/VIC10437/2020\|EPI_ISL_562910\|202 | .................... | .................... | .................... | .................... | .................... | .................... |
| K | 7692 | hCoV-19/Australia/VIC10438/2020\|EPI_ISL_562911\|202 | .................... | .................... | .................... | .................... | .................... | .................... |
| K | 7693 | hCoV-19/Australia/VIC10439/2020\|EPI_ISL_562912\|202 | .................... | .................... | .................... | .................... | .................... | .................... |
| K | 7694 | hCoV-19/Australia/VIC10440/2020\|EPI_ISL_562913\|202 | .................... | .................... | .................... | .................... | .................... | .................... |
| K | 7695 | hCoV-19/Australia/VIC10443/2020\|EPI_ISL_562914\|202 | .................... | .................... | .................... | .................... | .................... | .................... |
| K | 7696 | hCoV-19/Australia/VIC10444/2020\|EPI_ISL_562915\|202 | .................... | .................... | .................... | .................... | .................... | .................... |
| K | 7697 | hCoV-19/Australia/VIC10454/2020\|EPI_ISL_562916\|202 | .................... | .................... | .................... | .................... | .................... | .................... |
| K | 7698 | hCoV-19/Australia/VIC10455/2020\|EPI_ISL_562917\|202 | .................... | .................... | .................... | .................... | .................... | .................... |
| K | 7700 | hCoV-19/Australia/VIC10461/2020\|EPI_ISL_562919\|202 | .................... | .................... | .................... | .................... | .................... | .................... |
| K | 7701 | hCoV-19/Australia/VIC10463/2020\|EPI_ISL_562920\|202 | .................... | .................... | .................... | .................... | .................... | .................... |
| K | 7703 | hCoV-19/Australia/VIC10466/2020\|EPI_ISL_562922\|202 | .................... | .................... | .................... | .................... | .................... | .................... |
| K | 7704 | hCoV-19/Australia/VIC10467/2020\|EPI_ISL_562923\|202 | .................... | .................... | .................... | .................... | .................... | .................... |
| K | 7705 | hCoV-19/Australia/VIC10468/2020\|EPI_ISL_562924\|202 | .................... | .................... | .................... | .................... | .................... | .................... |
| K | 7706 | hCoV-19/Australia/VIC10471/2020\|EPI_ISL_562925\|202 | .................... | .................... | .................... | .................... | .................... | .................... |
| K | 7708 | hCoV-19/Australia/VIC10475/2020\|EPI_ISL_562927\|202 | .................... | .................... | .................... | .................... | .................... | .................... |
| K | 7709 | hCoV-19/Australia/VIC10477/2020\|EPI_ISL_562928\|202 | .................... | .................... | .................... | .................... | .................... | .................... |
| K | 7710 | hCoV-19/Australia/VIC10482/2020\|EPI_ISL_562929\|202 | .................... | .................... | .................... | .................... | .................... | .................... |
| K | 7711 | hCoV-19/Australia/VIC10485/2020\|EPI_ISL_562930\|202 | .................... | .................... | .................... | .................... | .................... | .................... |
| K | 7712 | hCoV-19/Australia/VIC10490/2020\|EPI_ISL_562931\|202 | .................... | .................... | .................... | .................... | .................... | .................... |
| K | 7713 | hCoV-19/Australia/VIC10491/2020\|EPI_ISL_562932\|202 | .................... | .................... | .................... | .................... | .................... | .................... |
| K | 7714 | hCoV-19/Australia/VIC10496/2020\|EPI_ISL_562933\|202 | .................... | .................... | .................... | .................... | .................... | .................... |
| K | 7715 | hCoV-19/Australia/VIC10497/2020\|EPI_ISL_562934\|202 | .................... | .................... | .................... | .................... | .................... | .................... |
| K | 7716 | hCoV-19/Australia/VIC10498/2020\|EPI_ISL_562935\|202 | .................... | .................... | .................... | .................... | .................... | .................... |
| K | 7717 | hCoV-19/Australia/VIC10499/2020\|EPI_ISL_562936\|202 | .................... | .................... | .................... | .................... | .................... | .................... |
| K | 7719 | hCoV-19/Australia/VIC10509/2020\|EPI_ISL_562938\|202 | .................... | .................... | .................... | .................... | .................... | .................... |
| K | 7720 | hCoV-19/Australia/VIC10511/2020\|EPI_ISL_562939\|202 | .................... | .................... | .................... | .................... | .................... | .................... |
| K | 7721 | hCoV-19/Australia/VIC10515/2020\|EPI_ISL_562940\|202 | .................... | .................... | .................... | .................... | .................... | .................... |
| K | 7722 | hCoV-19/Australia/VIC10516/2020\|EPI_ISL_562941\|202 | .................... | .................... | .................... | .................... | .................... | .................... |
| K | 7723 | hCoV-19/Australia/VIC10517/2020\|EPI_ISL_562942\|202 | .................... | .................... | .................... | .................... | .................... | .................... |
| K | 7724 | hCoV-19/Australia/VIC10520/2020\|EPI_ISL_562943\|202 | .................... | .................... | .................... | .................... | .................... | .................... |
| K | 7726 | hCoV-19/Australia/VIC10523/2020\|EPI_ISL_562945\|202 | .................... | .................... | .................... | .................... | .................... | .................... |
| K | 7727 | hCoV-19/Australia/VIC10525/2020\|EPI_ISL_562946\|202 | .................... | .................... | .................... | .................... | .................... | .................... |
| K | 7728 | hCoV-19/Australia/VIC10527/2020\|EPI_ISL_562947\|202 | .................... | .................... | .................... | .................... | .................... | .................... |

| | | | | | | | | |
|---|---|---|---|---|---|---|---|---|
| K | 7729 hCoV-19/Australia/VIC10530/2020\|EPI_ISL_562948\|202 | ···················· | ···················· | ····················· | ···················· | ···················· | ···················· | |
| K | 7730 hCoV-19/Australia/VIC10532/2020\|EPI_ISL_562949\|202 | ···················· | ···················· | ····················· | ···················· | ···················· | ···················· | |
| K | 7731 hCoV-19/Australia/VIC10535/2020\|EPI_ISL_562950\|202 | ···················· | ···················· | ····················· | ···················· | ···················· | ···················· | |
| K | 7732 hCoV-19/Australia/VIC10536/2020\|EPI_ISL_562951\|202 | ···················· | ···················· | ····················· | ···················· | ···················· | ··················C· | 1 |
| K | 7733 hCoV-19/Australia/VIC10538/2020\|EPI_ISL_562952\|202 | ···················· | ···················· | ····················· | ···················· | ···················· | ···················· | |
| K | 7734 hCoV-19/Australia/VIC10542/2020\|EPI_ISL_562953\|202 | ···················· | ···················· | ····················· | ···················· | ···················· | ···················· | |
| K | 7735 hCoV-19/Australia/VIC10548/2020\|EPI_ISL_562954\|202 | ···················· | ···················· | ····················· | ···················· | ···················· | ···················· | |
| K | 7736 hCoV-19/Australia/VIC10551/2020\|EPI_ISL_562955\|202 | ···················· | ···················· | ····················· | ···················· | ···················· | ···················· | |
| K | 7737 hCoV-19/Australia/VIC10554/2020\|EPI_ISL_562956\|202 | ···················· | ···················· | ····················· | ···················· | ···················· | ···················· | |
| K | 7738 hCoV-19/Australia/VIC10556/2020\|EPI_ISL_562957\|202 | ···················· | ···················· | ····················· | ···················· | ···················· | ···················· | |
| K | 7739 hCoV-19/Australia/VIC10562/2020\|EPI_ISL_562958\|202 | ···················· | ···················· | ····················· | ···················· | ···················· | ···················· | |
| K | 7740 hCoV-19/Australia/VIC10563/2020\|EPI_ISL_562959\|202 | ···················· | ···················· | ····················· | ···················· | ···················· | ···················· | |
| K | 7741 hCoV-19/Australia/VIC10564/2020\|EPI_ISL_562960\|202 | ···················· | ···················· | ····················· | ···················· | ···················· | ···················· | |
| K | 7742 hCoV-19/Australia/VIC10565/2020\|EPI_ISL_562961\|202 | ···················· | ···················· | ····················· | ···················· | ···················· | ···················· | |
| K | 7743 hCoV-19/Australia/VIC10569/2020\|EPI_ISL_562962\|202 | ···················· | ···················· | ····················· | ···················· | ···················· | ···················· | |
| K | 7744 hCoV-19/Australia/VIC10571/2020\|EPI_ISL_562963\|202 | ···················· | ···················· | ····················· | ···················· | ···················· | ···················· | |
| K | 7745 hCoV-19/Australia/VIC10574/2020\|EPI_ISL_562964\|202 | ···················· | ···················· | ····················· | ···················· | ···················· | ···················· | |
| K | 7746 hCoV-19/Australia/VIC10575/2020\|EPI_ISL_562965\|202 | ···················· | ···················· | ····················· | ···················· | ···················· | ···················· | |
| K | 7748 hCoV-19/Australia/VIC10579/2020\|EPI_ISL_562967\|202 | ···················· | ···················· | ····················· | ···················· | ···················· | ···················· | |
| K | 7749 hCoV-19/Australia/VIC10580/2020\|EPI_ISL_562968\|202 | ···················· | ···················· | ····················· | ···················· | ···················· | ···················· | |
| K | 7750 hCoV-19/Australia/VIC10581/2020\|EPI_ISL_562969\|202 | ···················· | ···················· | ····················· | ···················· | ···················· | ···················· | |
| K | 7751 hCoV-19/Australia/VIC10584/2020\|EPI_ISL_562970\|202 | ···················· | ···················· | ····················· | ···················· | ···················· | ···················· | |
| K | 7752 hCoV-19/Australia/VIC10585/2020\|EPI_ISL_562971\|202 | ···················· | ···················· | ····················· | ···················· | ···················· | ···················· | |
| K | 7753 hCoV-19/Australia/VIC10588/2020\|EPI_ISL_562972\|202 | ···················· | ···················· | ····················· | ···················· | ···················· | ···················· | |
| K | 7755 hCoV-19/Australia/VIC10590/2020\|EPI_ISL_562974\|202 | ···················· | ···················· | ····················· | ···················· | ···················· | ···················· | |
| K | 7756 hCoV-19/Australia/VIC10593/2020\|EPI_ISL_562975\|202 | ···················· | ···················· | ····················· | ···················· | ···················· | ···················· | |
| K | 7758 hCoV-19/Australia/VIC10595/2020\|EPI_ISL_562977\|202 | ···················· | ···················· | ····················· | ···················· | ···················· | ···················· | |
| K | 7760 hCoV-19/Australia/VIC10598/2020\|EPI_ISL_562979\|202 | ···················· | ···················· | ····················· | ···················· | ···················· | ···················· | |
| K | 7761 hCoV-19/Australia/VIC10599/2020\|EPI_ISL_562980\|202 | ···················· | ···················· | ····················· | ···················· | ···················· | ···················· | |
| K | 7763 hCoV-19/Australia/VIC8584/2020\|EPI_ISL_565100\|2020 | ···················· | ···················· | ····················· | ···················· | ···················· | ···················· | |
| K | 7764 hCoV-19/Australia/VIC8586/2020\|EPI_ISL_565101\|2020 | ···················· | ···················· | ····················· | ···················· | ···················· | ···················· | |
| K | 7765 hCoV-19/Australia/VIC8588/2020\|EPI_ISL_565102\|2020 | ···················· | ···················· | ····················· | ···················· | ···················· | ···················· | |
| K | 7766 hCoV-19/Australia/VIC8589/2020\|EPI_ISL_565103\|2020 | ···················· | ···················· | ····················· | ···················· | ···················· | ···················· | |
| K | 7767 hCoV-19/Australia/VIC8592/2020\|EPI_ISL_565104\|2020 | ···················· | ···················· | ····················· | ···················· | ···················· | ···················· | |
| K | 7768 hCoV-19/Australia/VIC8602/2020\|EPI_ISL_565105\|2020 | ···················· | ···················· | ····················· | ···················· | ···················· | ···················· | |
| K | 7769 hCoV-19/Australia/VIC8603/2020\|EPI_ISL_565106\|2020 | ···················· | ···················· | ····················· | ···················· | ···················· | ···················· | |
| K | 7770 hCoV-19/Australia/VIC8604/2020\|EPI_ISL_565107\|2020 | ···················· | ···················· | ····················· | ···················· | ···················· | ···················· | |
| K | 7771 hCoV-19/Australia/VIC8605/2020\|EPI_ISL_565108\|2020 | ···················· | ···················· | ····················· | ···················· | ···················· | ···················· | |
| K | 7772 hCoV-19/Australia/VIC8610/2020\|EPI_ISL_565109\|2020 | ···················· | ···················· | ····················· | ···················· | ···················· | ···················· | |
| K | 7773 hCoV-19/Australia/VIC8611/2020\|EPI_ISL_565110\|2020 | ···················· | ···················· | ····················· | ···················· | ···················· | ···················· | |
| K | 7774 hCoV-19/Australia/VIC8619/2020\|EPI_ISL_565111\|2020 | ···················· | ···················· | ····················· | ···················· | ···················· | ···················· | |
| K | 7775 hCoV-19/Australia/VIC8621/2020\|EPI_ISL_565112\|2020 | ···················· | ···················· | ····················· | ···················· | ···················· | ···················· | |
| K | 7776 hCoV-19/Australia/VIC8622/2020\|EPI_ISL_565113\|2020 | ···················· | ···················· | ····················· | ···················· | ···················· | ···················· | |
| K | 7777 hCoV-19/Australia/VIC8623/2020\|EPI_ISL_565114\|2020 | ···················· | ···················· | ····················· | ···················· | ···················· | ···················· | |
| K | 7778 hCoV-19/Australia/VIC8624/2020\|EPI_ISL_565115\|2020 | ···················· | ···················· | ····················· | ···················· | ···················· | ···················· | |
| K | 7779 hCoV-19/Australia/VIC8625/2020\|EPI_ISL_565116\|2020 | ···················· | ···················· | ····················· | ···················· | ···················· | ···················· | |
| K | 7780 hCoV-19/Australia/VIC8626/2020\|EPI_ISL_565117\|2020 | ···················· | ···················· | ····················· | ···················· | ···················· | ···················· | |
| K | 7781 hCoV-19/Australia/VIC8627/2020\|EPI_ISL_565118\|2020 | ···················· | ···················· | ····················· | ···················· | ···················· | ···················· | |
| K | 7782 hCoV-19/Australia/VIC8628/2020\|EPI_ISL_565119\|2020 | ···················· | ···················· | ····················· | ···················· | ···················· | ···················· | |
| K | 7783 hCoV-19/Australia/VIC8629/2020\|EPI_ISL_565120\|2020 | ···················· | ···················· | ····················· | ···················· | ···················· | ···················· | |
| K | 7784 hCoV-19/Australia/VIC8630/2020\|EPI_ISL_565121\|2020 | ···················· | ···················· | ····················· | ···················· | ···················· | ···················· | |
| K | 7785 hCoV-19/Australia/VIC8631/2020\|EPI_ISL_565122\|2020 | ···················· | ···················· | ····················· | ···················· | ···················· | ···················· | |
| K | 7786 hCoV-19/Australia/VIC8632/2020\|EPI_ISL_565123\|2020 | ···················· | ···················· | ····················· | ···················· | ···················· | ···················· | |
| K | 7787 hCoV-19/Australia/VIC8633/2020\|EPI_ISL_565124\|2020 | ···················· | ···················· | ····················· | ···················· | ···················· | ···················· | |
| K | 7788 hCoV-19/Australia/VIC8638/2020\|EPI_ISL_565125\|2020 | ···················· | ···················· | ····················· | ···················· | ···················· | ···················· | |
| K | 7789 hCoV-19/Australia/VIC8642/2020\|EPI_ISL_565126\|2020 | ···················· | ···················· | ····················· | ···················· | ···················· | ···················· | |
| K | 7790 hCoV-19/Australia/VIC8643/2020\|EPI_ISL_565127\|2020 | ···················· | ···················· | ····················· | ···················· | ···················· | ···················· | |
| K | 7791 hCoV-19/Australia/VIC8644/2020\|EPI_ISL_565128\|2020 | ···················· | ···················· | ····················· | ···················· | ···················· | ···················· | |
| K | 7792 hCoV-19/Australia/VIC8649/2020\|EPI_ISL_565129\|2020 | ···················· | ···················· | ····················· | ···················· | ···················· | ···················· | |
| K | 7793 hCoV-19/Australia/VIC8651/2020\|EPI_ISL_565130\|2020 | ···················· | ···················· | ····················· | ···················· | ···················· | ···················· | |
| K | 7794 hCoV-19/Australia/VIC8652/2020\|EPI_ISL_565131\|2020 | ···················· | ···················· | ····················· | ···················· | ···················· | ···················· | |
| K | 7795 hCoV-19/Australia/VIC8658/2020\|EPI_ISL_565132\|2020 | ···················· | ···················· | ····················· | ···················· | ···················· | ···················· | |
| K | 7796 hCoV-19/Australia/VIC8660/2020\|EPI_ISL_565133\|2020 | ···················· | ···················· | ····················· | ···················· | ···················· | ···················· | |
| K | 7797 hCoV-19/Australia/VIC8661/2020\|EPI_ISL_565134\|2020 | ···················· | ···················· | ····················· | ···················· | ···················· | ···················· | |
| K | 7798 hCoV-19/Australia/VIC8665/2020\|EPI_ISL_565135\|2020 | ···················· | ···················· | ····················· | ···················· | ···················· | ···················· | |
| K | 7799 hCoV-19/Australia/VIC8668/2020\|EPI_ISL_565136\|2020 | ···················· | ···················· | ···K················· | ···················· | ···················· | ····M··············· | |
| K | 7800 hCoV-19/Australia/VIC8670/2020\|EPI_ISL_565137\|2020 | ···················· | ···················· | ····················· | ···················· | ···················· | ···················· | |
| K | 7801 hCoV-19/Australia/VIC8673/2020\|EPI_ISL_565138\|2020 | ···················· | ···················· | ····················· | ···················· | ···················· | ···················· | |
| K | 7802 hCoV-19/Australia/VIC8674/2020\|EPI_ISL_565139\|2020 | ···················· | ···················· | ····················· | ···················· | ···················· | ···················· | |
| K | 7803 hCoV-19/Australia/VIC8678/2020\|EPI_ISL_565140\|2020 | ···················N | ···················· | ···K················· | ···················· | ···················· | ···················· | 1 |
| K | 7804 hCoV-19/Australia/VIC8691/2020\|EPI_ISL_565141\|2020 | ···················· | ···················· | ····················· | ···················· | ···················· | ···················· | |
| K | 7805 hCoV-19/Australia/VIC8692/2020\|EPI_ISL_565142\|2020 | ···················· | ···················· | ···K················· | ···················· | ···················· | ···················· | |
| K | 7806 hCoV-19/Australia/VIC8698/2020\|EPI_ISL_565143\|2020 | ···················· | ···················· | ····················· | ···················· | ···················· | ···················· | |
| K | 7807 hCoV-19/Australia/VIC8701/2020\|EPI_ISL_565144\|2020 | ···················· | ···················· | ····················· | ···················· | ···················· | ···················· | |
| K | 7808 hCoV-19/Australia/VIC8702/2020\|EPI_ISL_565145\|2020 | ···················· | ···················· | ····················· | ···················· | ···················· | ···················· | |
| K | 7809 hCoV-19/Australia/VIC8704/2020\|EPI_ISL_565146\|2020 | ···················· | ···················· | ····················· | ···················· | ···················· | ···················· | |
| K | 7810 hCoV-19/Australia/VIC8705/2020\|EPI_ISL_565147\|2020 | ···················· | ···················· | ····················· | ···················· | ···················· | ···················· | |
| K | 7811 hCoV-19/Australia/VIC8708/2020\|EPI_ISL_565148\|2020 | ···················· | ···················· | ····················· | ···················· | ···················· | ···················· | |
| K | 7812 hCoV-19/Australia/VIC8709/2020\|EPI_ISL_565149\|2020 | ···················· | ···················· | ····················· | ···················· | ···················· | ···················· | |
| K | 7813 hCoV-19/Australia/VIC8710/2020\|EPI_ISL_565150\|2020 | ···················· | ···················· | ····················· | ···················· | ···················· | ···················· | |

| | | | | | | | |
|---|---|---|---|---|---|---|---|
| K | 7814 hCoV-19/Australia/VIC8711/2020\|EPI_ISL_565151\|2020 | ···················· | ···················· | ····················· | ···················· | ···················· | ···················· |
| K | 7815 hCoV-19/Australia/VIC8712/2020\|EPI_ISL_565152\|2020 | ···················· | ···················· | ····················· | ···················· | ···················· | ···················· |
| K | 7816 hCoV-19/Australia/VIC8715/2020\|EPI_ISL_565153\|2020 | ···················· | ···················· | ····················· | ···················· | ···················· | ···················· |
| K | 7817 hCoV-19/Australia/VIC8716/2020\|EPI_ISL_565154\|2020 | ···················· | ···················· | ····················· | ···················· | ···················· | ···················· |
| K | 7819 hCoV-19/Australia/VIC8720/2020\|EPI_ISL_565156\|2020 | ···················· | ···················· | ····················· | ···················· | ···················· | ···················· |
| K | 7820 hCoV-19/Australia/VIC8721/2020\|EPI_ISL_565157\|2020 | ···················· | ···················· | ····················· | ···················· | ···················· | ···················· |
| K | 7821 hCoV-19/Australia/VIC8728/2020\|EPI_ISL_565158\|2020 | ···················· | ···················· | ····················· | ···················· | ···················· | ···················· |
| K | 7822 hCoV-19/Australia/VIC8729/2020\|EPI_ISL_565159\|2020 | ···················· | ···················· | ····················· | ···················· | ···················· | ···················· |
| K | 7824 hCoV-19/Australia/VIC8761/2020\|EPI_ISL_565181\|2020 | ···················· | ···················· | ····················· | ···················· | ···················· | ···················· |
| K | 7825 hCoV-19/Australia/VIC8767/2020\|EPI_ISL_565182\|2020 | ···················· | ···················· | ····················· | ···················· | ···················· | ···················· |
| K | 7826 hCoV-19/Australia/VIC8768/2020\|EPI_ISL_565183\|2020 | ···················· | ···················· | ····················· | ···················· | ···················· | ···················· |
| K | 7827 hCoV-19/Australia/VIC8769/2020\|EPI_ISL_565184\|2020 | ···················· | ···················· | ····················· | ···················· | ···················· | ···················· |
| K | 7828 hCoV-19/Australia/VIC8773/2020\|EPI_ISL_565185\|2020 | ···················· | ···················· | ····················· | ···················· | ···················· | ···················· |
| K | 7829 hCoV-19/Australia/VIC8774/2020\|EPI_ISL_565186\|2020 | ···················· | ···················· | ····················· | ···················· | ···················· | ···················· |
| K | 7830 hCoV-19/Australia/VIC8776/2020\|EPI_ISL_565187\|2020 | ···················· | ···················· | ····················· | ···················· | ···················· | ···················· |
| K | 7832 hCoV-19/Australia/VIC8779/2020\|EPI_ISL_565189\|2020 | ···················· | ···················· | ····················· | ···················· | ···················· | ···················· |
| K | 7833 hCoV-19/Australia/VIC8781/2020\|EPI_ISL_565190\|2020 | ···················· | ···················· | ····················· | ···················· | ···················· | ···················· |
| K | 7834 hCoV-19/Australia/VIC8782/2020\|EPI_ISL_565191\|2020 | ···················· | ···················· | ····················· | ···················· | N··N················ | ···················· |
| K | 7835 hCoV-19/Australia/VIC8783/2020\|EPI_ISL_565192\|2020 | ···················· | ···················· | ····················· | ···················· | ···················· | ···················· |
| K | 7836 hCoV-19/Australia/VIC8784/2020\|EPI_ISL_565193\|2020 | ···················· | ···················· | ····················· | ···················· | ···················· | ···················· |
| K | 7837 hCoV-19/Australia/VIC8785/2020\|EPI_ISL_565194\|2020 | ···················· | ···················· | ····················· | ···················· | ···················· | ···················· |
| K | 7839 hCoV-19/Australia/VIC8789/2020\|EPI_ISL_565196\|2020 | ···················· | ···················· | ····················· | ···················· | ···················· | ···················· |
| K | 7840 hCoV-19/Australia/VIC8790/2020\|EPI_ISL_565197\|2020 | ···················· | ···················· | ····················· | ···················· | ···················· | ···················· |
| K | 7841 hCoV-19/Australia/VIC8791/2020\|EPI_ISL_565198\|2020 | ···················· | ···················· | ····················· | ···················· | ···················· | ···················· |
| K | 7842 hCoV-19/Australia/VIC8793/2020\|EPI_ISL_565199\|2020 | ···················· | ···················· | ····················· | ···················· | ···················· | ···················· |
| K | 7843 hCoV-19/England/MILK-9C2C85/2020\|EPI_ISL_566425\|20 | ···················· | ···················· | ····················· | ···················· | ···················· | ···················· |
| K | 7844 hCoV-19/Wales/CAMC-9B1CB6/2020\|EPI_ISL_566429\|2020 | ···················· | ···················· | ····················· | ···················· | ···················· | ···················· |
| K | 7845 hCoV-19/England/MILK-9C21A4/2020\|EPI_ISL_566430\|20 | ···················· | ·A·················· | ····················· | ···················· | ···················· | ··A················· |
| K | 7846 hCoV-19/England/MILK-9C2308/2020\|EPI_ISL_566431\|20 | ···················· | ···················· | ····················· | ···················· | ···················· | ···················· |
| K | 7847 hCoV-19/England/MILK-9C1F9C/2020\|EPI_ISL_566432\|20 | ···················· | ···················· | ····················· | ···················· | ···················· | ···················· |
| K | 7848 hCoV-19/England/MILK-9C2C0D/2020\|EPI_ISL_566433\|20 | ···················· | ···················· | ····················· | ···················· | ···················· | ···················· |
| K | 7849 hCoV-19/Wales/MILK-9C1FC9/2020\|EPI_ISL_566434\|2020 | ···················· | ···················· | ····················· | ···················· | ···················· | ···················· |
| K | 7850 hCoV-19/England/MILK-9C204D/2020\|EPI_ISL_566436\|20 | ···················· | ···················· | ····················· | ···················· | ···················· | ···················· |
| K | 7851 hCoV-19/England/MILK-9C2D37/2020\|EPI_ISL_566437\|20 | ···················· | ···················· | ····················· | ···················· | ···················· | ···················· |
| K | 7852 hCoV-19/England/MILK-9C212C/2020\|EPI_ISL_566438\|20 | ···················· | ···················· | ····················· | ···················· | ···················· | ···················· |
| K | 7853 hCoV-19/England/MILK-9C26D2/2020\|EPI_ISL_566439\|20 | ···················· | ···················· | ····················· | ···················· | ···················· | ···················· |
| K | 7854 hCoV-19/Wales/MILK-9C2EE9/2020\|EPI_ISL_566440\|2020 | ···················· | ···················· | ····················· | ···················· | ···················· | ···················· |
| K | 7855 hCoV-19/England/MILK-9C2793/2020\|EPI_ISL_566441\|20 | ···················· | ···················· | ····················· | ···················· | ···················· | ···················· |
| K | 7856 hCoV-19/England/MILK-9C2881/2020\|EPI_ISL_566443\|20 | ···················· | ···················· | ····················· | ···················· | ···················· | ···················· |
| K | 7857 hCoV-19/England/CAMC-9B22C2/2020\|EPI_ISL_566444\|20 | ···················· | ···················· | ····················· | ···················· | ···················· | ···················· |
| K | 7858 hCoV-19/England/CAMC-9B1BD7/2020\|EPI_ISL_566445\|20 | ···················· | ···················· | ····················· | ···················· | ······T············· | ···················· |
| K | 7859 hCoV-19/England/MILK-9C210E/2020\|EPI_ISL_566446\|20 | ···················· | ···················· | ····················· | ···················· | ···················· | ···················· |
| K | 7860 hCoV-19/Wales/MILK-9C2362/2020\|EPI_ISL_566447\|2020 | ···················· | ···················· | ····················· | ···················· | ···················· | ···················· |
| K | 7861 hCoV-19/England/MILK-9C22A1/2020\|EPI_ISL_566448\|20 | ···················· | ···················· | ····················· | ···················· | ···················· | ···················· |
| K | 7862 hCoV-19/England/MILK-9C2827/2020\|EPI_ISL_566449\|20 | ···················· | ···················· | ····················· | ···················· | ······T············· | ···················· |
| K | 7863 hCoV-19/England/MILK-9C2C3A/2020\|EPI_ISL_566450\|20 | ···················· | ·A·················· | ····················· | ···················· | ···················· | ··A················· |
| K | 7864 hCoV-19/England/MILK-9C1E71/2020\|EPI_ISL_566451\|20 | ···················· | ···················· | ····················· | ···················· | ···················· | ···················· |
| K | 7865 hCoV-19/England/MILK-9C2687/2020\|EPI_ISL_566452\|20 | ···················· | ···················· | ····················· | ···················· | ···················· | ···················· |
| K | 7866 hCoV-19/England/CAMC-9B1CA7/2020\|EPI_ISL_566453\|20 | ···················· | ···················· | ····················· | ···················· | ···················· | ···················· |
| K | 7867 hCoV-19/England/MILK-9C239F/2020\|EPI_ISL_566454\|20 | ···················· | ···················· | ····················· | ···················· | ···················· | ···················· |
| K | 7868 hCoV-19/England/CAMC-9B1B8C/2020\|EPI_ISL_566457\|20 | ···················· | ···················· | ····················· | ···················· | ···················· | ···················· |
| K | 7869 hCoV-19/England/MILK-9C1E35/2020\|EPI_ISL_566458\|20 | ···················· | ···················· | ····················· | ···················· | ···················· | ···················· |
| K | 7870 hCoV-19/England/MILK-9C2C67/2020\|EPI_ISL_566459\|20 | ···················· | ·A·······A·········· | ····················· | ···················· | ···················· | ···················· |
| K | 7871 hCoV-19/England/MILK-9C24BA/2020\|EPI_ISL_566460\|20 | ···················· | ···················· | ····················· | ···················· | ···················· | ···················· |
| K | 7872 hCoV-19/England/MILK-9C2A30/2020\|EPI_ISL_566461\|20 | ···················· | ···················· | ····················· | ···················· | ···················· | ···················· |
| K | 7873 hCoV-19/England/MILK-9C2872/2020\|EPI_ISL_566462\|20 | ···················· | ···················· | ····················· | ···················· | ···················· | ···················· |
| K | 7874 hCoV-19/England/MILK-9C2DCE/2020\|EPI_ISL_566463\|20 | ···················· | ···················· | ····················· | ···················· | ···················· | ···················· |
| K | 7875 hCoV-19/England/MILK-9C2353/2020\|EPI_ISL_566465\|20 | ···················· | ···················· | ····················· | ···················· | ···················· | ···················· |
| K | 7876 hCoV-19/England/MILK-9C272A/2020\|EPI_ISL_566466\|20 | ···················· | ···················· | ····················· | ···················· | ···················· | ···················· |
| K | 7877 hCoV-19/England/CAMC-9B1B13/2020\|EPI_ISL_566467\|20 | ···················· | ···················· | ····················· | ···················· | ···················· | ···················· |
| K | 7878 hCoV-19/England/MILK-9C2890/2020\|EPI_ISL_566468\|20 | ···················· | ···················· | ····················· | ···················· | ···················· | ···················· |
| K | 7879 hCoV-19/Wales/MILK-9C203E/2020\|EPI_ISL_566469\|2020 | ···················· | ···················· | ····················· | ···················· | ···················· | ···················· |
| K | 7880 hCoV-19/England/MILK-9C1EAE/2020\|EPI_ISL_566470\|20 | ···················· | ···················· | ····················· | ···················· | ···················· | ···················· |
| K | 7881 hCoV-19/Wales/MILK-9C1E53/2020\|EPI_ISL_566471\|2020 | ···················· | ···················· | ····················· | ···················· | ···················· | ···················· |
| K | 7882 hCoV-19/England/MILK-9C29CA/2020\|EPI_ISL_566472\|20 | ···················· | ···················· | ····················· | ···················· | ···················· | ···················· |
| K | 7883 hCoV-19/England/MILK-9C2F22/2020\|EPI_ISL_566475\|20 | ···················· | ·A·················· | ····················· | ···················· | ···················· | ··A················· |
| K | 7884 hCoV-19/England/MILK-9C2CA3/2020\|EPI_ISL_566477\|20 | ···················· | ···················· | ····················· | ···················· | ···················· | ···················· |
| K | 7885 hCoV-19/England/MILK-9C2766/2020\|EPI_ISL_566478\|20 | ···················· | ···················· | ····················· | ···················· | ···················· | ···················· |
| K | 7886 hCoV-19/Wales/MILK-9C1F41/2020\|EPI_ISL_566480\|2020 | ···················· | ···················· | ····················· | ···················· | ···················· | ···················· |
| K | 7887 hCoV-19/England/MILK-9C2AA9/2020\|EPI_ISL_566481\|20 | ···················· | ···················· | ····················· | ···················· | ···················· | ···················· |
| K | 7888 hCoV-19/England/MILK-9C2E70/2020\|EPI_ISL_566482\|20 | ···················· | ···················· | ····················· | ···················· | ···················· | ···················· |
| K | 7889 hCoV-19/England/MILK-9C2EBC/2020\|EPI_ISL_566483\|20 | ···················· | ···················· | ····················· | ···················· | ···················· | ···················· |
| K | 7890 hCoV-19/England/MILK-9C270C/2020\|EPI_ISL_566484\|20 | ···················· | ···················· | ····················· | ···················· | ···················· | ···················· |
| K | 7891 hCoV-19/England/MILK-9C28DC/2020\|EPI_ISL_566485\|20 | ···················· | ···················· | ····················· | ···················· | ···················· | ···················· |
| K | 7892 hCoV-19/England/MILK-9C2E8F/2020\|EPI_ISL_566487\|20 | ···················· | ···················· | ····················· | ···················· | ···················· | ···················· |
| K | 7893 hCoV-19/England/MILK-9C2B6A/2020\|EPI_ISL_566488\|20 | ···················· | ···················· | ····················· | ···················· | ···················· | ···················· |
| K | 7894 hCoV-19/Wales/MILK-9C1E44/2020\|EPI_ISL_566489\|2020 | ···················· | ···················· | ····················· | ···················· | ···················· | ···················· |
| K | 7895 hCoV-19/England/ALDP-9C6270/2020\|EPI_ISL_567756\|20 | ···················· | ···················· | ····················· | ···················· | ···················· | ···················· |
| K | 7896 hCoV-19/England/ALDP-9C5CEC/2020\|EPI_ISL_567757\|20 | ···················· | ···················· | ····················· | ···················· | ···················· | ···················· |
| K | 7897 hCoV-19/England/ALDP-9C5C55/2020\|EPI_ISL_567758\|20 | ···················· | ·A·················· | ····················· | ···················· | ···················· | ··A················· |

| | | | | | | | |
|---|---|---|---|---|---|---|---|
| K | 7898 hCoV-19/England/ALDP-9C4EF6/2020\|EPI_ISL_567760\|20 | ···················· | ···················· | ···················· | ···················· | ···················· | ···················· |
| K | 7899 hCoV-19/England/ALDP-9C4C56/2020\|EPI_ISL_567761\|20 | ···················· | ···················· | ···················· | ···················· | ···················· | ···················· |
| K | 7900 hCoV-19/England/ALDP-9C4BE0/2020\|EPI_ISL_567762\|20 | ···················· | ·A·················· | ···················· | ···················· | ···················· | ··A················· |
| K | 7901 hCoV-19/England/ALDP-9C5262/2020\|EPI_ISL_567763\|20 | ···················· | ···················· | ···················· | ···················· | ···················· | ···················· |
| K | 7902 hCoV-19/England/ALDP-9C547B/2020\|EPI_ISL_567764\|20 | ···················· | ···················· | ···················· | ···················· | ···················· | ···················· |
| K | 7903 hCoV-19/England/ALDP-9C6182/2020\|EPI_ISL_567766\|20 | ···················· | ···················· | ···················· | ···················· | ···················· | ···················· |
| K | 7904 hCoV-19/England/ALDP-9C6058/2020\|EPI_ISL_567768\|20 | ···················· | ···················· | ···················· | ···················· | ···················· | ···················· |
| K | 7905 hCoV-19/England/ALDP-9C4F3F/2020\|EPI_ISL_567769\|20 | ···················· | ···················· | ···················· | ···················· | ···················· | ···················· |
| K | 7906 hCoV-19/England/ALDP-9C5280/2020\|EPI_ISL_567771\|20 | ···················· | ···················· | ···················· | ···················· | ···················· | ···················· |
| K | 7907 hCoV-19/England/ALDP-9C4EC9/2020\|EPI_ISL_567772\|20 | ···················· | ···················· | ···················· | ···················· | ···················· | ···················· |
| K | 7908 hCoV-19/England/ALDP-9C52AE/2020\|EPI_ISL_567773\|20 | ···················· | ···················· | ···················· | ···················· | ···················· | ···················· |
| K | 7909 hCoV-19/England/ALDP-9C5059/2020\|EPI_ISL_567774\|20 | ···················· | ···················· | ···················· | ···················· | ···················· | ···················· |
| K | 7910 hCoV-19/England/ALDP-9C5D25/2020\|EPI_ISL_567775\|20 | ···················· | ·A·······A·········· | ···················· | ···················· | ···················· | ···················· |
| K | 7911 hCoV-19/England/ALDP-9C603A/2020\|EPI_ISL_567776\|20 | ···················· | ···················· | ···················· | ···················· | ···················· | ···················· |
| K | 7912 hCoV-19/England/ALDP-9C550F/2020\|EPI_ISL_567777\|20 | ···················· | ···················· | ···················· | ···················· | ···················· | ···················· |
| K | 7913 hCoV-19/England/ALDP-9C4F99/2020\|EPI_ISL_567778\|20 | ···················· | ···················· | ···················· | ···················· | ···················· | ···················· |
| K | 7914 hCoV-19/England/ALDP-9C636E/2020\|EPI_ISL_567779\|20 | ···················· | ·A·················· | ···················· | ···················· | ···················· | ··A················· |
| K | 7915 hCoV-19/England/ALDP-9C4E05/2020\|EPI_ISL_567780\|20 | ···················· | ···················· | ···················· | ···················· | ···················· | ···················· |
| K | 7916 hCoV-19/England/ALDP-9C4DEA/2020\|EPI_ISL_567781\|20 | ···················· | ···················· | ···················· | ···················· | ···················· | ···················· |
| K | 7917 hCoV-19/Wales/ALDP-9C5772/2020\|EPI_ISL_567782\|2020 | ···················· | ···················· | ···················· | ···················· | ···················· | ···················· |
| K | 7918 hCoV-19/England/ALDP-9C594F/2020\|EPI_ISL_567783\|20 | ···················· | ···················· | ···················· | ···················· | ···················· | ···················· |
| K | 7919 hCoV-19/England/ALDP-9C61CE/2020\|EPI_ISL_567784\|20 | ···················· | ···················· | ···················· | ···················· | ···················· | ···················· |
| K | 7920 hCoV-19/England/ALDP-9C6207/2020\|EPI_ISL_567785\|20 | ···················· | ···················· | ···················· | ···················· | ···················· | ···················· |
| K | 7921 hCoV-19/England/ALDP-9C55A5/2020\|EPI_ISL_567786\|20 | ···················· | ···················· | ···················· | ···················· | ···················· | ···················· |
| K | 7922 hCoV-19/England/ALDP-9C5E9B/2020\|EPI_ISL_567787\|20 | ···················· | ·A·················· | ···················· | ···················· | ···················· | ··A················· |
| K | 7923 hCoV-19/England/ALDP-9C5C91/2020\|EPI_ISL_567788\|20 | ···················· | ···················· | ···················· | ···················· | ···················· | ···················· |
| K | 7924 hCoV-19/England/ALDP-9C4B68/2020\|EPI_ISL_567789\|20 | ···················· | ···················· | ···················· | ···················· | ···················· | ···················· |
| K | 7925 hCoV-19/England/ALDP-9C4D53/2020\|EPI_ISL_567790\|20 | ···················· | ···················· | ···················· | ···················· | ···················· | ···················· |
| K | 7926 hCoV-19/England/ALDP-9C57EB/2020\|EPI_ISL_567791\|20 | ···················· | ···················· | ···················· | ···················· | ···················· | ···················· |
| K | 7927 hCoV-19/Wales/ALDP-9C5138/2020\|EPI_ISL_567792\|2020 | ···················· | ···················· | ···················· | ···················· | ······T············· | ···················· |
| K | 7928 hCoV-19/England/ALDP-9C6322/2020\|EPI_ISL_567793\|20 | ···················· | ···················· | ···················· | ···················· | ···················· | ···················· |
| K | 7929 hCoV-19/England/ALDP-9C5736/2020\|EPI_ISL_567794\|20 | ···················· | ···················· | ···················· | ···················· | ···················· | ···················· |
| K | 7930 hCoV-19/England/ALDP-9C56DF/2020\|EPI_ISL_567795\|20 | ···················· | ···················· | ···················· | ···················· | ····T··············· | ···················· |
| K | 7931 hCoV-19/England/ALDP-9C5350/2020\|EPI_ISL_567796\|20 | ···················· | ···················· | ···················· | ···················· | ···················· | ···················· |
| K | 7932 hCoV-19/England/ALDP-9C5402/2020\|EPI_ISL_567797\|20 | ···················· | ·A·················· | ···················· | ···················· | ···················· | ··A················· |
| K | 7933 hCoV-19/England/ALDP-9C5CFB/2020\|EPI_ISL_567798\|20 | ···················· | ···················· | ···················· | ···················· | ···················· | ···················· |
| K | 7934 hCoV-19/England/ALDP-9C548A/2020\|EPI_ISL_567799\|20 | ···················· | ···················· | ···················· | ···················· | ···················· | ···················· |
| K | 7935 hCoV-19/England/ALDP-9BE150/2020\|EPI_ISL_567901\|20 | ···················· | ···················· | ···················· | ···················· | ····T··············· | ···················· |
| K | 7936 hCoV-19/England/ALDP-9BDBAE/2020\|EPI_ISL_567902\|20 | ···················· | ···················· | ···················· | ···················· | ···················· | ···················· |
| K | 7937 hCoV-19/USA/NM-UNM-00610/2020\|EPI_ISL_560325\|2020- | ···················· | ···················· | ···················· | ···················· | ···················· | ···················· |
| K | 7938 hCoV-19/USA/NM-UNM-00683/2020\|EPI_ISL_560326\|2020- | ···················· | ···················· | ···················· | ···················· | ···················· | ···················· |
| K | 7939 hCoV-19/USA/NM-UNM-00659/2020\|EPI_ISL_560330\|2020- | ···················· | ···················· | ···················· | ···················· | ···················· | ···················· |
| K | 7940 hCoV-19/USA/NM-UNM-00678/2020\|EPI_ISL_560332\|2020- | ···················· | ···················· | ···················· | ···················· | ···················· | ···················· |
| K | 7941 hCoV-19/USA/NM-UNM-00660/2020\|EPI_ISL_560333\|2020- | ···················· | ···················· | ···················· | ···················· | ···················· | ···················· |
| K | 7942 hCoV-19/USA/NM-UNM-00646/2020\|EPI_ISL_560334\|2020- | ···················· | ···················· | ···················· | ···················· | ···················· | ···················· |
| K | 7943 hCoV-19/USA/NM-UNM-00692/2020\|EPI_ISL_560336\|2020- | ···················· | ···················· | ···················· | ···················· | ···················· | ···················· |
| K | 7944 hCoV-19/USA/NM-UNM-00670/2020\|EPI_ISL_560338\|2020- | ···················· | ···················· | ···················· | ···················· | ···················· | ···················· |
| K | 7945 hCoV-19/USA/NM-UNM-00684/2020\|EPI_ISL_560340\|2020- | ···················· | ···················· | ···················· | ···················· | ···················· | ···················· |
| K | 7946 hCoV-19/USA/NM-UNM-00609/2020\|EPI_ISL_560347\|2020- | ···················· | ···················· | ···················· | ···················· | ···················· | ···················· |
| K | 7947 hCoV-19/USA/NM-UNM-00616/2020\|EPI_ISL_560348\|2020- | ···················· | ···················· | ···················· | ···················· | ···················· | ···················· |
| K | 7948 hCoV-19/USA/NM-UNM-00633/2020\|EPI_ISL_560350\|2020- | ···················· | ···················· | ···················· | ···················· | ···················· | ···················· |
| K | 7949 hCoV-19/USA/NM-UNM-00599/2020\|EPI_ISL_560354\|2020- | ···················· | ···················· | ···················· | ···················· | ···················· | ···················· |
| K | 7950 hCoV-19/Botswana/BOT0455/2020\|EPI_ISL_560386\|2020- | ···················· | ···················· | ···················· | ···················· | ···················· | ···················· |
| K | 7951 hCoV-19/Lithuania/C20-05-R10/2020\|EPI_ISL_560391\|2 | ···················· | ···················· | ···················· | ···················· | ···················· | ···················· |
| K | 7952 hCoV-19/Lithuania/C20-05-R11/2020\|EPI_ISL_560392\|2 | ···················· | ···················· | ···················· | ···················· | ···················· | ···················· |
| K | 7953 hCoV-19/Lithuania/C20-05-R13/2020\|EPI_ISL_560393\|2 | ···················· | ···················· | ···················· | ···················· | ···················· | ···················· |
| K | 7954 hCoV-19/Lithuania/C20-05-R15/2020\|EPI_ISL_560394\|2 | ···················· | ···················· | ···················· | ···················· | ···················· | ···················· |
| K | 7955 hCoV-19/Lithuania/C20-05-R16/2020\|EPI_ISL_560395\|2 | ···················· | ···················· | ···················· | ···················· | ···················· | ···················· |
| K | 7956 hCoV-19/Lithuania/C20-05-R18/2020\|EPI_ISL_560396\|2 | ···················· | ···················· | ···················· | ···················· | ···················· | ···················· |
| K | 7957 hCoV-19/Lithuania/C20-05-R19/2020\|EPI_ISL_560397\|2 | ···················· | ···················· | ···················· | ···················· | ···················· | ···················· |
| K | 7958 hCoV-19/Lithuania/C20-05-R3/2020\|EPI_ISL_560399\|20 | ···················· | ···················· | ···················· | ···················· | ···················· | ···················· |
| K | 7959 hCoV-19/Australia/VIC10601/2020\|EPI_ISL_562982\|202 | ···················· | ···················· | ···················· | ···················· | ·········K·········· | ···················· |
| K | 7960 hCoV-19/Australia/VIC10603/2020\|EPI_ISL_562983\|202 | ···················· | ···················· | ···················· | ···················· | ···················· | ···················· |
| K | 7962 hCoV-19/Australia/VIC10605/2020\|EPI_ISL_562985\|202 | ···················· | ···················· | ···················· | ···················· | ···················· | ···················· |
| K | 7964 hCoV-19/Australia/VIC10609/2020\|EPI_ISL_562987\|202 | ···················· | ···················· | ···················· | ···················· | ···················· | ···················· |
| K | 7965 hCoV-19/Australia/VIC10610/2020\|EPI_ISL_562988\|202 | ···················· | ···················· | ···················· | ···················· | ···················· | ···················· |
| K | 7966 hCoV-19/Australia/VIC10611/2020\|EPI_ISL_562989\|202 | ···················· | ···················· | ···················· | ···················· | ···················· | ···················· |
| K | 7969 hCoV-19/Australia/VIC10615/2020\|EPI_ISL_562992\|202 | ···················· | ···················· | ···················· | ···················· | ···················· | ···················· |
| K | 7970 hCoV-19/Australia/VIC10616/2020\|EPI_ISL_562993\|202 | ···················· | ···················· | ···················· | ···················· | ···················· | ···················· |
| K | 7971 hCoV-19/Australia/VIC10618/2020\|EPI_ISL_562994\|202 | ···················· | ···················· | ···················· | ···················· | ···················· | ···················· |
| K | 7974 hCoV-19/Australia/VIC10622/2020\|EPI_ISL_562997\|202 | ···················· | ···················· | ···················· | ···················· | ···················· | ···················· |
| K | 7977 hCoV-19/Australia/VIC8732/2020\|EPI_ISL_565160\|2020 | ···················· | ···················· | ···················· | ···················· | ···················· | ···················· |
| K | 7978 hCoV-19/Australia/VIC8733/2020\|EPI_ISL_565161\|2020 | ···················· | ···················· | ···················· | ···················· | ···················· | ···················· |
| K | 7979 hCoV-19/Australia/VIC8735/2020\|EPI_ISL_565162\|2020 | ···················· | ···················· | ···················· | ···················· | ···················· | ···················· |
| K | 7980 hCoV-19/Australia/VIC8736/2020\|EPI_ISL_565163\|2020 | ···················· | ···················· | ···················· | ···················· | ···················· | ···················· |
| K | 7981 hCoV-19/Australia/VIC8737/2020\|EPI_ISL_565164\|2020 | ···················· | ···················· | ···················· | ···················· | ···················· | ···················· |
| K | 7982 hCoV-19/Australia/VIC8738/2020\|EPI_ISL_565165\|2020 | ···················· | ···················· | ···················· | ···················· | ···················· | ···················· |
| K | 7983 hCoV-19/Australia/VIC8739/2020\|EPI_ISL_565166\|2020 | ···················· | ···················· | ···················· | ···················· | ···················· | ···················· |
| K | 7984 hCoV-19/Australia/VIC8740/2020\|EPI_ISL_565167\|2020 | ········Y··········· | ···················· | ···················· | ···················· | ···················· | ···················· |
| K | 7985 hCoV-19/Australia/VIC8741/2020\|EPI_ISL_565168\|2020 | ···················· | ···················· | ···················· | ···················· | ···················· | ···················· |

| | | | | | | | | |
|---|---|---|---|---|---|---|---|---|
| K | 7986 hCoV-19/Australia/VIC8742/2020\|EPI_ISL_565169\|2020 | ···················· | ···················· | ···················· | ···················· | ···················· | ···················· | |
| K | 7987 hCoV-19/Australia/VIC8743/2020\|EPI_ISL_565170\|2020 | ···················· | ···················· | ···················· | ···················· | ···················· | ···················· | |
| K | 7988 hCoV-19/Australia/VIC8744/2020\|EPI_ISL_565171\|2020 | ···················· | ···················· | ···················· | ···················· | ···················· | ···················· | |
| K | 7989 hCoV-19/Australia/VIC8749/2020\|EPI_ISL_565172\|2020 | ···················· | ···················· | ···················· | ···················· | ···················· | ···················· | |
| K | 7990 hCoV-19/Australia/VIC8750/2020\|EPI_ISL_565173\|2020 | ···················· | ···················· | ···················· | ···················· | ···················· | ···················· | |
| K | 7991 hCoV-19/Australia/VIC8751/2020\|EPI_ISL_565174\|2020 | ···················· | ···················· | ···················· | ···················· | ···················· | ···················· | |
| K | 7992 hCoV-19/Australia/VIC8752/2020\|EPI_ISL_565175\|2020 | ···················· | ···················· | ···················· | ···················· | ···················· | ···················· | |
| K | 7993 hCoV-19/Australia/VIC8755/2020\|EPI_ISL_565176\|2020 | ···················· | ···················· | ···················· | ···················· | ···················· | ···················· | |
| K | 7994 hCoV-19/Australia/VIC8756/2020\|EPI_ISL_565177\|2020 | ···················· | ···················· | ···················· | ···················· | ···················· | ···················· | |
| K | 7995 hCoV-19/Australia/VIC8757/2020\|EPI_ISL_565178\|2020 | ···················· | ···················· | ···················· | ···················· | ···················· | ···················· | |
| K | 7996 hCoV-19/Australia/VIC8758/2020\|EPI_ISL_565179\|2020 | ···················· | ···················· | ···················· | ···················· | ···················· | ···················· | |
| K | 7997 hCoV-19/Australia/VIC8794/2020\|EPI_ISL_565200\|2020 | ···················· | ···················· | ···················· | ···················· | ···················· | ···················· | |
| K | 7998 hCoV-19/Australia/VIC8795/2020\|EPI_ISL_565201\|2020 | ···················· | ···················· | ···················· | ···················· | ···················· | ···················· | |
| K | 7999 hCoV-19/Australia/VIC8798/2020\|EPI_ISL_565202\|2020 | ···················· | ···················· | ···················· | ···················· | ···················· | ···················· | |
| K | 8000 hCoV-19/Australia/VIC8800/2020\|EPI_ISL_565203\|2020 | ···················· | ···················· | ···················· | ···················· | ···················· | ···················· | |
| K | 8001 hCoV-19/Australia/VIC8801/2020\|EPI_ISL_565204\|2020 | ···················· | ···················· | ···················· | ···················· | ···················· | ···················· | |
| K | 8002 hCoV-19/Australia/VIC8802/2020\|EPI_ISL_565205\|2020 | ···················· | ···················· | ···················· | ···················· | ···················· | ···················· | |
| K | 8004 hCoV-19/Australia/VIC8805/2020\|EPI_ISL_565207\|2020 | ···················· | ···M················ | ···················· | ···················· | ···················· | ···················· | |
| K | 8005 hCoV-19/Australia/VIC8806/2020\|EPI_ISL_565208\|2020 | ···················· | ···················· | ···················· | ···················· | NNN················· | ···················· | 1 |
| K | 8006 hCoV-19/Australia/VIC8807/2020\|EPI_ISL_565209\|2020 | ···················· | ···················· | ···················· | ···················· | ···················· | ···················· | |
| K | 8007 hCoV-19/Australia/VIC8811/2020\|EPI_ISL_565210\|2020 | ···················· | ···················· | ···················· | ···················· | ···················· | ···················· | |
| K | 8008 hCoV-19/Australia/VIC8813/2020\|EPI_ISL_565211\|2020 | ···················· | ···················· | ···················· | ···················· | ···················· | ···················· | |
| K | 8009 hCoV-19/Australia/VIC8814/2020\|EPI_ISL_565212\|2020 | ···················· | ···················· | ···················· | ···················· | ···················· | ···················· | |
| K | 8010 hCoV-19/Australia/VIC8815/2020\|EPI_ISL_565213\|2020 | ···················· | ···················· | ···················· | ···················· | ·······Y············ | ···················· | |
| K | 8011 hCoV-19/Australia/VIC8816/2020\|EPI_ISL_565214\|2020 | ···················· | ···················· | ···················· | ···················· | ···················· | ···················· | |
| K | 8012 hCoV-19/England/MILK-9C2520/2020\|EPI_ISL_566490\|20 | ···················· | ···················· | ···················· | ···················· | ···················· | ···················· | |
| K | 8013 hCoV-19/England/MILK-9C2863/2020\|EPI_ISL_566491\|20 | ···················· | ···················· | ···················· | ···················· | ···················· | ···················· | |
| K | 8014 hCoV-19/England/CAMC-9B1ADA/2020\|EPI_ISL_566493\|20 | ···················· | ···················· | ···················· | ···················· | ···················· | ···················· | |
| K | 8015 hCoV-19/England/CAMC-9B21E3/2020\|EPI_ISL_566495\|20 | ···················· | ···················· | ···················· | ···················· | ···················· | ···················· | |
| K | 8016 hCoV-19/England/MILK-9C2CEF/2020\|EPI_ISL_566496\|20 | ···················· | ···················· | ···················· | ···················· | ···················· | ···················· | |
| K | 8017 hCoV-19/England/MILK-9C2A12/2020\|EPI_ISL_566497\|20 | ···················· | ·A·················· | ···················· | ···················· | ···················· | ···A················ | |
| K | 8018 hCoV-19/England/MILK-9C2B97/2020\|EPI_ISL_566498\|20 | ···················· | ···················· | ···················· | ···················· | ···················· | ···················· | |
| K | 8019 hCoV-19/England/MILK-9C2784/2020\|EPI_ISL_566499\|20 | ···················· | ···················· | ···················· | ···················· | ···················· | ···················· | |
| K | 8020 hCoV-19/England/CAMC-9B1D2C/2020\|EPI_ISL_566500\|20 | ···················· | ···················· | ···················· | ···················· | ···················· | ···················· | |
| K | 8021 hCoV-19/Wales/MILK-9C2FB9/2020\|EPI_ISL_566503\|2020 | ···················· | ···················· | ···················· | ···················· | ···················· | ···················· | |
| K | 8022 hCoV-19/England/MILK-9C2B88/2020\|EPI_ISL_566506\|20 | ···················· | ·A·················· | ···················· | ···················· | ···················· | ···A················ | |
| K | 8023 hCoV-19/England/MILK-9C2159/2020\|EPI_ISL_566507\|20 | ···················· | ···················· | ···················· | ···················· | ···················· | ···················· | |
| K | 8024 hCoV-19/Wales/CAMC-9B1BB9/2020\|EPI_ISL_566508\|2020 | ···················· | ···················· | ···················· | ···················· | ···················· | ···················· | |
| K | 8025 hCoV-19/England/MILK-9C1F7E/2020\|EPI_ISL_566509\|20 | ···················· | ···················· | ···················· | ···················· | ···················· | ···················· | |
| K | 8026 hCoV-19/England/MILK-9C2229/2020\|EPI_ISL_566510\|20 | ···················· | ···················· | ···················· | ···················· | ···················· | ···················· | |
| K | 8027 hCoV-19/England/MILK-9C29BB/2020\|EPI_ISL_566511\|20 | ···················· | ···················· | ···················· | ···················· | ···················· | ···················· | |
| K | 8028 hCoV-19/England/MILK-9C2678/2020\|EPI_ISL_566512\|20 | ···················· | ···················· | ···················· | ···················· | ···················· | ···················· | |
| K | 8029 hCoV-19/England/MILK-9C1E17/2020\|EPI_ISL_566514\|20 | ···················· | ···················· | ···················· | ···················· | ···················· | ···················· | |
| K | 8030 hCoV-19/England/MILK-9C2CB2/2020\|EPI_ISL_566515\|20 | ···················· | ···················· | ···················· | ···················· | ···················· | ···················· | |
| K | 8031 hCoV-19/Wales/MILK-9C2EF8/2020\|EPI_ISL_566516\|2020 | ···················· | ···················· | ···················· | ···················· | ···················· | ···················· | |
| K | 8032 hCoV-19/England/MILK-9C2177/2020\|EPI_ISL_566517\|20 | ···················· | ···················· | ···················· | ···················· | ···················· | ···················· | |
| K | 8033 hCoV-19/England/CAMC-9B1BF5/2020\|EPI_ISL_566518\|20 | ···················· | ···················· | ···················· | ···················· | ···················· | ···················· | |
| K | 8034 hCoV-19/England/CAMC-9B1D1D/2020\|EPI_ISL_566519\|20 | ···················· | ···················· | ···················· | ···················· | ···················· | ···················· | |
| K | 8035 hCoV-19/Wales/MILK-9C1E62/2020\|EPI_ISL_566520\|2020 | ···················· | ···················· | ···················· | ···················· | ···················· | ···················· | |
| K | 8036 hCoV-19/England/MILK-9C1ECC/2020\|EPI_ISL_566521\|20 | ···················· | ···················· | ···················· | ···················· | ···················· | ···················· | |
| K | 8037 hCoV-19/England/MILK-9C2CD0/2020\|EPI_ISL_566522\|20 | ···················· | ···················· | ···················· | ···················· | ···················· | ···················· | |
| K | 8038 hCoV-19/England/MILK-9C25E4/2020\|EPI_ISL_566523\|20 | ···················· | ···················· | ···················· | ···················· | ···················· | ···················· | |
| K | 8039 hCoV-19/England/CAMC-9B25C9/2020\|EPI_ISL_566524\|20 | ···················· | ···················· | ···················· | ···················· | ···················· | ···················· | |
| K | 8040 hCoV-19/England/MILK-9C2BB5/2020\|EPI_ISL_566525\|20 | ···················· | ·A·················· | ···················· | ···················· | ···················· | ···A················ | |
| K | 8041 hCoV-19/England/MILK-9C299D/2020\|EPI_ISL_566526\|20 | ···················· | ···················· | ···················· | ···················· | ···················· | ···················· | |
| K | 8042 hCoV-19/England/MILK-9C2292/2020\|EPI_ISL_566527\|20 | ···················· | ···················· | ···················· | ···················· | ···················· | ···················· | |
| K | 8043 hCoV-19/England/CAMC-9B1C6B/2020\|EPI_ISL_566528\|20 | ···················· | ·A·················· | ···················· | ···················· | ···················· | ···A················ | |
| K | 8044 hCoV-19/England/MILK-9C297F/2020\|EPI_ISL_566530\|20 | ···················· | ···················· | ···················· | ···················· | ···················· | ···················· | |
| K | 8045 hCoV-19/England/MILK-9C24D8/2020\|EPI_ISL_566531\|20 | ···················· | ···················· | ···················· | ···················· | ···················· | ···················· | |
| K | 8046 hCoV-19/England/MILK-9C2BC4/2020\|EPI_ISL_566532\|20 | ···················· | ·A·················· | ···················· | ···················· | ···················· | ···A················ | |
| K | 8047 hCoV-19/England/MILK-9C211D/2020\|EPI_ISL_566533\|20 | ···················· | ···················· | ···················· | ···················· | ···················· | ···················· | |
| K | 8048 hCoV-19/England/CAMC-9B1B5F/2020\|EPI_ISL_566534\|20 | ···················· | ···················· | ···················· | ···················· | ···················· | ···················· | |
| K | 8049 hCoV-19/England/MILK-9C2960/2020\|EPI_ISL_566535\|20 | ···················· | ···················· | ···················· | ···················· | ···················· | ···················· | |
| K | 8050 hCoV-19/England/MILK-9C2BF1/2020\|EPI_ISL_566536\|20 | ···················· | ···················· | ···················· | ···················· | ···················· | ···················· | |
| K | 8051 hCoV-19/England/MILK-9C2D19/2020\|EPI_ISL_566539\|20 | ···················· | ···················· | ···················· | ···················· | ···················· | ···················· | |
| K | 8052 hCoV-19/England/MILK-9C206B/2020\|EPI_ISL_566540\|20 | ···················· | ·A·················· | ···················· | ···················· | ···················· | ···A················ | |
| K | 8053 hCoV-19/Wales/MILK-9C20B6/2020\|EPI_ISL_566541\|2020 | ···················· | ···················· | ···················· | ···················· | ···················· | ···················· | |
| K | 8054 hCoV-19/England/MILK-9C1F6F/2020\|EPI_ISL_566542\|20 | ···················· | ···················· | ···················· | ···················· | ···················· | ···················· | |
| K | 8055 hCoV-19/England/CAMC-9B1D77/2020\|EPI_ISL_566543\|20 | ···················· | ·A·················· | ···················· | ···················· | ···················· | ···A················ | |
| K | 8056 hCoV-19/England/CAMC-9B1AE9/2020\|EPI_ISL_566544\|20 | ···················· | ···················· | ···················· | ···················· | ···················· | ···················· | |
| K | 8057 hCoV-19/England/MILK-9C2238/2020\|EPI_ISL_566545\|20 | ···················· | ···················· | ···················· | ···················· | ···················· | ···················· | |
| K | 8058 hCoV-19/England/ALDP-9C5569/2020\|EPI_ISL_567800\|20 | ···················· | ···················· | ···················· | ···················· | ···················· | ···················· | |
| K | 8059 hCoV-19/England/ALDP-9C53D8/2020\|EPI_ISL_567801\|20 | ···················· | ···················· | ···················· | ···················· | ···················· | ···················· | |
| K | 8060 hCoV-19/England/ALDP-9C5499/2020\|EPI_ISL_567802\|20 | ···················· | ···················· | ···················· | ···················· | ···················· | ···················· | |
| K | 8061 hCoV-19/England/ALDP-9C4B86/2020\|EPI_ISL_567804\|20 | ···················· | ···················· | ···················· | ···················· | ···················· | ···················· | |
| K | 8062 hCoV-19/England/ALDP-9C5BD0/2020\|EPI_ISL_567805\|20 | ···················· | ···················· | ···················· | ···················· | ···················· | ···················· | |
| K | 8063 hCoV-19/England/ALDP-9C4D71/2020\|EPI_ISL_567806\|20 | ···················· | ···················· | ···················· | ···················· | ···················· | ···················· | |
| K | 8064 hCoV-19/England/ALDP-9C5095/2020\|EPI_ISL_567807\|20 | ···················· | ···················· | ···················· | ···················· | ···················· | ···················· | |
| K | 8065 hCoV-19/Wales/ALDP-9C5EE6/2020\|EPI_ISL_567808\|2020 | ···················· | ···················· | ···················· | ···················· | ······T············· | ···················· | |
| K | 8066 hCoV-19/England/ALDP-9C5323/2020\|EPI_ISL_567809\|20 | ···················· | ·A·················· | ···················· | ···················· | ···················· | ···A················ | |

| | | | | | | | | |
|---|---|---|---|---|---|---|---|---|
| K | 8067 | hCoV-19/England/ALDP-9C5DAD/2020\|EPI_ISL_567810\|20 | ···················· | ·A·················· | ···················· | ···················· | ···················· | ··A················· |
| K | 8068 | hCoV-19/England/ALDP-9C4F6C/2020\|EPI_ISL_567813\|20 | ···················· | ···················· | ···················· | ···················· | ···················· | ···················· |
| K | 8069 | hCoV-19/England/ALDP-9C501D/2020\|EPI_ISL_567814\|20 | ···················· | ···················· | ···················· | ···················· | ···················· | ···················· |
| K | 8070 | hCoV-19/England/ALDP-9C61BF/2020\|EPI_ISL_567815\|20 | ···················· | ···················· | ···················· | ···················· | ···················· | ···················· |
| K | 8071 | hCoV-19/England/ALDP-9C5DBC/2020\|EPI_ISL_567816\|20 | ···················· | ···················· | ···················· | ···················· | ···················· | ···················· |
| K | 8072 | hCoV-19/England/ALDP-9C54D5/2020\|EPI_ISL_567817\|20 | ···················· | ···················· | ···················· | ···················· | ···················· | ···················· |
| K | 8073 | hCoV-19/England/ALDP-9C5F7A/2020\|EPI_ISL_567818\|20 | ···················· | ···················· | ···················· | ···················· | ···················· | ···················· |
| K | 8074 | hCoV-19/England/ALDP-9C5D16/2020\|EPI_ISL_567819\|20 | ···················· | ···················· | ···················· | ···················· | ···················· | ···················· |
| K | 8075 | hCoV-19/England/ALDP-9C58AC/2020\|EPI_ISL_567820\|20 | ···················· | ···················· | ···················· | ···················· | ···················· | ···················· |
| K | 8076 | hCoV-19/England/ALDP-9C5C46/2020\|EPI_ISL_567821\|20 | ···················· | ···················· | ···················· | ···················· | ···················· | ···················· |
| K | 8077 | hCoV-19/England/ALDP-9C638C/2020\|EPI_ISL_567822\|20 | ···················· | ···················· | ···················· | ···················· | ···················· | ···················· |
| K | 8078 | hCoV-19/England/ALDP-9C5FC5/2020\|EPI_ISL_567823\|20 | ···················· | ·A·················· | ···················· | ···················· | ···················· | ··A················· |
| K | 8079 | hCoV-19/England/ALDP-9C595E/2020\|EPI_ISL_567824\|20 | ···················· | ···················· | ···················· | ···················· | ···················· | ···················· |
| K | 8080 | hCoV-19/England/ALDP-9C4BFF/2020\|EPI_ISL_567825\|20 | ···················· | ···················· | ···················· | ···················· | ···················· | ···················· |
| K | 8081 | hCoV-19/England/ALDP-9C5E04/2020\|EPI_ISL_567826\|20 | ···················· | ·A·················· | ···················· | ···················· | ···················· | ··A················· |
| K | 8082 | hCoV-19/England/ALDP-9C4E9C/2020\|EPI_ISL_567827\|20 | ···················· | ···················· | ···················· | ···················· | ···················· | ···················· |
| K | 8083 | hCoV-19/England/ALDP-9C5411/2020\|EPI_ISL_567828\|20 | ···················· | ···················· | ···················· | ···················· | ···················· | ···················· |
| K | 8084 | hCoV-19/England/ALDP-9C642F/2020\|EPI_ISL_567829\|20 | ···················· | ···················· | ···················· | ···················· | ···················· | ···················· |
| K | 8085 | hCoV-19/England/ALDP-9C4BA4/2020\|EPI_ISL_567830\|20 | ···················· | ···················· | ···················· | ···················· | ···················· | ···················· |
| K | 8086 | hCoV-19/England/ALDP-9C5244/2020\|EPI_ISL_567832\|20 | ···················· | ···················· | ···················· | ···················· | ···················· | ···················· |
| K | 8087 | hCoV-19/England/ALDP-9C4F02/2020\|EPI_ISL_567834\|20 | ···················· | ···················· | ···················· | ···················· | ···················· | ···················· |
| K | 8088 | hCoV-19/England/ALDP-9C4DBD/2020\|EPI_ISL_567835\|20 | ···················· | ·A·················· | ···················· | ···················· | ···················· | ···················· |
| K | 8089 | hCoV-19/England/ALDP-9C545D/2020\|EPI_ISL_567836\|20 | ···················· | ···················· | ···················· | ···················· | ···················· | ···················· |
| K | 8090 | hCoV-19/England/ALDP-9C5253/2020\|EPI_ISL_567837\|20 | ···················· | ···················· | ···················· | ···················· | ···················· | ···················· |
| K | 8091 | hCoV-19/England/ALDP-9C4C0B/2020\|EPI_ISL_567838\|20 | ···················· | ···················· | ···················· | ···················· | ···················· | ···················· |
| K | 8092 | hCoV-19/England/ALDP-9C4C38/2020\|EPI_ISL_567839\|20 | ···················· | ···················· | ···················· | ···················· | ···················· | ···················· |
| K | 8093 | hCoV-19/England/ALDP-9C51ED/2020\|EPI_ISL_567840\|20 | ···················· | ···················· | ···················· | ···················· | ···················· | ···················· |
| K | 8094 | hCoV-19/England/ALDP-9C589D/2020\|EPI_ISL_567841\|20 | ···················· | ···················· | ···················· | ···················· | ···················· | ···················· |
| K | 8095 | hCoV-19/England/ALDP-9C4E14/2020\|EPI_ISL_567842\|20 | ···················· | ···················· | ···················· | ···················· | ···················· | ···················· |
| K | 8096 | hCoV-19/England/ALDP-9C5314/2020\|EPI_ISL_567843\|20 | ···················· | ·A·················· | ···················· | ···················· | ···················· | ··A················· |
| K | 8097 | hCoV-19/England/ALDP-9C5930/2020\|EPI_ISL_567844\|20 | ···················· | ···················· | ···················· | ···················· | ···················· | ···················· |
| K | 8098 | hCoV-19/England/ALDP-9C5587/2020\|EPI_ISL_567846\|20 | ···················· | ···················· | ···················· | ···················· | ···················· | ···················· |
| K | 8099 | hCoV-19/England/ALDP-9C5332/2020\|EPI_ISL_567847\|20 | ···················· | ···················· | ···················· | ···················· | ···················· | ···················· |
| K | 8100 | hCoV-19/England/ALDP-9C5A00/2020\|EPI_ISL_567848\|20 | ···················· | ···················· | ···················· | ···················· | ···················· | ···················· |
| K | 8101 | hCoV-19/England/ALDP-9C4E6F/2020\|EPI_ISL_567849\|20 | ···················· | ···················· | ···················· | ···················· | ···················· | ···················· |
| K | 8102 | hCoV-19/England/ALDP-9C4ED8/2020\|EPI_ISL_567850\|20 | ···················· | ···················· | ···················· | ···················· | ···················· | ···················· |
| K | 8103 | hCoV-19/England/ALDP-9C552D/2020\|EPI_ISL_567851\|20 | ···················· | ···················· | ···················· | ···················· | ···················· | ···················· |
| K | 8104 | hCoV-19/England/ALDP-9C4D9F/2020\|EPI_ISL_567852\|20 | ···················· | ···················· | ···················· | ···················· | ···················· | ···················· |
| K | 8105 | hCoV-19/England/ALDP-9C57DC/2020\|EPI_ISL_567853\|20 | ···················· | ···················· | ···················· | ···················· | ···················· | ···················· |
| K | 8106 | hCoV-19/England/ALDP-9C4D17/2020\|EPI_ISL_567854\|20 | ···················· | ·A·················· | ···················· | ···················· | ···················· | ··A················· |
| K | 8107 | hCoV-19/England/ALDP-9C5068/2020\|EPI_ISL_567855\|20 | ···················· | ···················· | ···················· | ···················· | ···················· | ···················· |
| K | 8108 | hCoV-19/England/ALDP-9C5727/2020\|EPI_ISL_567856\|20 | ···················· | ···················· | ···················· | ···················· | ···················· | ···················· |
| K | 8109 | hCoV-19/England/ALDP-9C5DE9/2020\|EPI_ISL_567857\|20 | ···················· | ·A·················· | ···················· | ···················· | ···················· | ··A················· |
| K | 8110 | hCoV-19/England/ALDP-9C4F7B/2020\|EPI_ISL_567858\|20 | ···················· | ···················· | ···················· | ···················· | ···················· | ···················· |
| K | 8111 | hCoV-19/England/ALDP-9C58CA/2020\|EPI_ISL_567861\|20 | ···················· | ···················· | ···················· | ···················· | ···················· | ···················· |
| K | 8112 | hCoV-19/England/ALDP-9C63E6/2020\|EPI_ISL_567863\|20 | ···················· | ···················· | ···················· | ···················· | ···················· | ···················· |
| K | 8113 | hCoV-19/England/ALDP-9C56B1/2020\|EPI_ISL_567864\|20 | ···················· | ···················· | ···················· | ···················· | ···················· | ···················· |
| K | 8114 | hCoV-19/England/ALDP-9C5657/2020\|EPI_ISL_567865\|20 | ···················· | ···················· | ···················· | ···················· | ···················· | ···················· |
| K | 8115 | hCoV-19/England/ALDP-9C4D35/2020\|EPI_ISL_567866\|20 | ···················· | ···················· | ···················· | ···················· | ···················· | ···················· |
| K | 8116 | hCoV-19/England/ALDP-9C6049/2020\|EPI_ISL_567867\|20 | ···················· | ···················· | ···················· | ···················· | ···················· | ···················· |
| K | 8117 | hCoV-19/England/ALDP-9C57CD/2020\|EPI_ISL_567868\|20 | ···················· | ···················· | ···················· | ···················· | ···················· | ···················· |
| K | 8118 | hCoV-19/England/ALDP-9C599A/2020\|EPI_ISL_567869\|20 | ···················· | ···················· | ···················· | ···················· | ···················· | ···················· |
| K | 8119 | hCoV-19/England/ALDP-9C5D43/2020\|EPI_ISL_567870\|20 | ···················· | ···················· | ···················· | ···················· | ···················· | ···················· |
| K | 8120 | hCoV-19/England/ALDP-9C5EC8/2020\|EPI_ISL_567871\|20 | ···················· | ···················· | ···················· | ···················· | ···················· | ···················· |
| K | 8121 | hCoV-19/England/ALDP-9C5165/2020\|EPI_ISL_567872\|20 | ···················· | ···················· | ···················· | ···················· | ···················· | ···················· |
| K | 8122 | hCoV-19/England/ALDP-9C61FB/2020\|EPI_ISL_567873\|20 | ···················· | ···················· | ···················· | ···················· | ···················· | ···················· |
| K | 8123 | hCoV-19/England/ALDP-9C63B9/2020\|EPI_ISL_567874\|20 | ···················· | ···················· | ···················· | ···················· | ···················· | ···················· |
| K | 8124 | hCoV-19/England/ALDP-9C539C/2020\|EPI_ISL_567875\|20 | ···················· | ···················· | ···················· | ···················· | ···················· | ···················· |
| K | 8125 | hCoV-19/England/ALDP-9C4FF3/2020\|EPI_ISL_567876\|20 | ···················· | ···················· | ···················· | ···················· | ···················· | ···················· |
| K | 8126 | hCoV-19/USA/WA-UW-21746/2020\|EPI_ISL_571000\|2020-0 | ···················· | ···················· | ···················· | ···················· | ···················· | ···················· |
| K | 8127 | hCoV-19/USA/WA-UW-21752/2020\|EPI_ISL_571001\|2020-0 | ···················· | ···················· | ···················· | ···················· | ···················· | ···················· |
| K | 8128 | hCoV-19/USA/WA-UW-21753/2020\|EPI_ISL_571002\|2020-0 | ···················· | ···················· | ···················· | ···················· | ···················· | ···················· |
| K | 8129 | hCoV-19/USA/WA-UW-21756/2020\|EPI_ISL_571003\|2020-0 | ···················· | ···················· | ···················· | ···················· | ···················· | ···················· |
| K | 8130 | hCoV-19/USA/WA-UW-21771/2020\|EPI_ISL_571004\|2020-0 | ···················· | ···················· | ···················· | ···················· | ···················· | ···················· |
| K | 8131 | hCoV-19/USA/WA-UW-21784/2020\|EPI_ISL_571005\|2020-0 | ···················· | ···················· | ···················· | ···················· | ···················· | ···················· |
| K | 8132 | hCoV-19/USA/WA-UW-21786/2020\|EPI_ISL_571006\|2020-0 | ···················· | ·A·················· | ···················· | ···················· | ···················· | ···················· |
| K | 8133 | hCoV-19/USA/WA-UW-21960/2020\|EPI_ISL_571007\|2020-0 | ···················· | ···················· | ···················· | ···················· | ···················· | ···················· |
| K | 8134 | hCoV-19/USA/WA-UW-21985/2020\|EPI_ISL_571008\|2020-0 | ···················· | ···················· | ···················· | ···················· | ···················· | ···················· |
| K | 8135 | hCoV-19/USA/WA-UW-21986/2020\|EPI_ISL_571009\|2020-0 | ···················· | ···················· | ···················· | ···················· | ···················· | ···················· |
| K | 8136 | hCoV-19/USA/WA-UW-21987/2020\|EPI_ISL_571010\|2020-0 | ···················· | ···················· | ···················· | ···················· | ···················· | ···················· |
| K | 8137 | hCoV-19/USA/WA-UW-21998/2020\|EPI_ISL_571011\|2020-0 | ···················· | ·A·················· | ···················· | ···················· | ···················· | ···················· |
| K | 8138 | hCoV-19/USA/WA-UW-22003/2020\|EPI_ISL_571012\|2020-0 | ···················· | ···················· | ···················· | ···················· | ···················· | ···················· |
| K | 8139 | hCoV-19/USA/WA-UW-22016/2020\|EPI_ISL_571013\|2020-0 | ···················· | ···················· | ···················· | ···················· | ···················· | ···················· |
| K | 8140 | hCoV-19/USA/WA-UW-22038/2020\|EPI_ISL_571014\|2020-0 | ···················· | ···················· | ···················· | ···················· | ···················· | ···················· |
| K | 8141 | hCoV-19/USA/WA-UW-22039/2020\|EPI_ISL_571015\|2020-0 | ···················· | ···················· | ···················· | ···················· | ···················· | ···················· |
| L | 1 | hCoV-19/USA/CA-QDX-1035/2020\|EPI_ISL_571700\|2020-0 | ···················· | ···················· | ···················· | ···················· | ···················· | ···················· |
| L | 2 | hCoV-19/USA/CA-QDX-1036/2020\|EPI_ISL_571701\|2020-0 | ···················· | ···················· | ···················· | ···················· | ···················· | ···················· |
| L | 3 | hCoV-19/USA/CA-QDX-1037/2020\|EPI_ISL_571702\|2020-0 | ···················· | ···················· | ···················· | ···················· | ···················· | ···················· |
| L | 4 | hCoV-19/USA/CA-QDX-1038/2020\|EPI_ISL_571703\|2020-0 | ···················· | ···················· | ···················· | ···················· | ···················· | ···················· |
| L | 5 | hCoV-19/USA/CA-QDX-1039/2020\|EPI_ISL_571704\|2020-0 | ···················· | ···················· | ···················· | ···················· | ···················· | ···················· |

| | | | | | | | | |
|---|---|---|---|---|---|---|---|---|
| L | 6 | hCoV-19/USA/CA-QDX-1040/2020\|EPI_ISL_571705\|2020-0 | ···················· | ···················· | ····················· | ···················· | ···················· | ···················· |
| L | 7 | hCoV-19/USA/CA-QDX-1041/2020\|EPI_ISL_571706\|2020-0 | ···················· | ···················· | ····················· | ···················· | ···················· | ···················· |
| L | 8 | hCoV-19/USA/CA-QDX-1042/2020\|EPI_ISL_571707\|2020-0 | ···················· | ···················· | ····················· | ···················· | ···················· | ···················· |
| L | 9 | hCoV-19/USA/CA-QDX-1043/2020\|EPI_ISL_571708\|2020-0 | ···················· | ···················· | ····················· | ···················· | ···················· | ···················· |
| L | 10 | hCoV-19/USA/CA-QDX-1044/2020\|EPI_ISL_571709\|2020-0 | ···················· | ···················· | ····················· | ···················· | ···················· | ···················· |
| L | 11 | hCoV-19/USA/CA-QDX-1045/2020\|EPI_ISL_571710\|2020-0 | ···················· | ···················· | ····················· | ···················· | ···················· | ···················· |
| L | 12 | hCoV-19/USA/CA-QDX-1049/2020\|EPI_ISL_571711\|2020-0 | ···················· | ···················· | ····················· | ···················· | ···················· | ···················· |
| L | 13 | hCoV-19/USA/CA-QDX-1051/2020\|EPI_ISL_571712\|2020-0 | ···················· | ···················· | ····················· | ···················· | ···················· | ···················· |
| L | 14 | hCoV-19/USA/CA-QDX-1053/2020\|EPI_ISL_571713\|2020-0 | ···················· | ···················· | ····················· | ···················· | ···················· | ···················· |
| L | 15 | hCoV-19/USA/CA-QDX-1057/2020\|EPI_ISL_571714\|2020-0 | ···················· | ···················· | ····················· | ···················· | ···················· | ···················· |
| L | 16 | hCoV-19/USA/CA-QDX-1059/2020\|EPI_ISL_571715\|2020-0 | ···················· | ···················· | ····················· | ···················· | ···················· | ···················· |
| L | 17 | hCoV-19/USA/CO-QDX-1055/2020\|EPI_ISL_571716\|2020-0 | ···················· | ···················· | ····················· | ···················· | ···················· | ···················· |
| L | 18 | hCoV-19/USA/IN-QDX-1004/2020\|EPI_ISL_571717\|2020-0 | ···················· | ···················· | ····················· | ···················· | ···················· | ···················· |
| L | 19 | hCoV-19/USA/IN-QDX-951/2020\|EPI_ISL_571718\|2020-03 | ···················· | ···················· | ····················· | ···················· | ···················· | ···················· |
| L | 20 | hCoV-19/USA/IN-QDX-963/2020\|EPI_ISL_571719\|2020-03 | ···················· | ···················· | ····················· | ···················· | ···················· | ···················· |
| L | 21 | hCoV-19/USA/IN-QDX-964/2020\|EPI_ISL_571720\|2020-03 | ···················· | ···················· | ····················· | ···················· | ···················· | ···················· |
| L | 22 | hCoV-19/USA/IN-QDX-993/2020\|EPI_ISL_571721\|2020-03 | ···················· | ···················· | ····················· | ···················· | ···················· | ···················· |
| L | 23 | hCoV-19/USA/MI-QDX-1046/2020\|EPI_ISL_571722\|2020-0 | ···················· | ···················· | ····················· | ···················· | ···················· | ···················· |
| L | 24 | hCoV-19/USA/MI-QDX-1047/2020\|EPI_ISL_571723\|2020-0 | ···················· | ···················· | ····················· | ···················· | ···················· | ···················· |
| L | 25 | hCoV-19/USA/MI-QDX-1048/2020\|EPI_ISL_571724\|2020-0 | ···················· | ···················· | ····················· | ···················· | ···················· | ···················· |
| L | 26 | hCoV-19/USA/MI-QDX-1050/2020\|EPI_ISL_571725\|2020-0 | ···················· | ···················· | ····················· | ···················· | ···················· | ···················· |
| L | 27 | hCoV-19/USA/MI-QDX-1052/2020\|EPI_ISL_571726\|2020-0 | ···················· | ···················· | ····················· | ···················· | ···················· | ···················· |
| L | 28 | hCoV-19/USA/MN-QDX-1000/2020\|EPI_ISL_571727\|2020-0 | ···················· | ···················· | ····················· | ···················· | ···················· | ···················· |
| L | 29 | hCoV-19/USA/MN-QDX-1001/2020\|EPI_ISL_571728\|2020-0 | ···················· | ···················· | ····················· | ···················· | ···················· | ···················· |
| L | 30 | hCoV-19/USA/MN-QDX-1002/2020\|EPI_ISL_571729\|2020-0 | ···················· | ···················· | ····················· | ···················· | ···················· | ···················· |
| L | 31 | hCoV-19/USA/MN-QDX-1003/2020\|EPI_ISL_571730\|2020-0 | ···················· | ···················· | ····················· | ···················· | ···················· | ···················· |
| L | 32 | hCoV-19/USA/MN-QDX-1005/2020\|EPI_ISL_571731\|2020-0 | ···················· | ···················· | ····················· | ···················· | ···················· | ···················· |
| L | 33 | hCoV-19/USA/MN-QDX-1006/2020\|EPI_ISL_571732\|2020-0 | ···················· | ···················· | ····················· | ···················· | ···················· | ···················· |
| L | 34 | hCoV-19/USA/MN-QDX-1007/2020\|EPI_ISL_571733\|2020-0 | ···················· | ···················· | ····················· | ···················· | ···················· | ···················· |
| L | 35 | hCoV-19/USA/MN-QDX-1008/2020\|EPI_ISL_571734\|2020-0 | ···················· | ···················· | ····················· | ···················· | ···················· | ···················· |
| L | 36 | hCoV-19/USA/MN-QDX-1009/2020\|EPI_ISL_571735\|2020-0 | ···················· | ···················· | ····················· | ···················· | ···················· | ···················· |
| L | 37 | hCoV-19/USA/MN-QDX-1010/2020\|EPI_ISL_571736\|2020-0 | ···················· | ···················· | ····················· | ···················· | ···················· | ···················· |
| L | 38 | hCoV-19/USA/MN-QDX-1011/2020\|EPI_ISL_571737\|2020-0 | ···················· | ···················· | ····················· | ···················· | ···················· | ···················· |
| L | 39 | hCoV-19/USA/MN-QDX-1012/2020\|EPI_ISL_571738\|2020-0 | ···················· | ···················· | ····················· | ···················· | ···················· | ···················· |
| L | 40 | hCoV-19/USA/MN-QDX-1013/2020\|EPI_ISL_571739\|2020-0 | ···················· | ···················· | ····················· | ···················· | ···················· | ···················· |
| L | 41 | hCoV-19/USA/MN-QDX-1014/2020\|EPI_ISL_571740\|2020-0 | ···················· | ···················· | ····················· | ···················· | ···················· | ···················· |
| L | 42 | hCoV-19/USA/MN-QDX-1015/2020\|EPI_ISL_571741\|2020-0 | ···················· | ···················· | ····················· | ···················· | ···················· | ···················· |
| L | 43 | hCoV-19/USA/MN-QDX-1016/2020\|EPI_ISL_571742\|2020-0 | ···················· | ···················· | ····················· | ···················· | ···················· | ···················· |
| L | 44 | hCoV-19/USA/MN-QDX-1017/2020\|EPI_ISL_571743\|2020-0 | ···················· | ···················· | ····················· | ···················· | ···················· | ···················· |
| L | 45 | hCoV-19/USA/MN-QDX-1018/2020\|EPI_ISL_571744\|2020-0 | ···················· | ···················· | ····················· | ···················· | ···················· | ···················· |
| L | 46 | hCoV-19/USA/MN-QDX-1019/2020\|EPI_ISL_571745\|2020-0 | ···················· | ···················· | ····················· | ···················· | ···················· | ···················· |
| L | 47 | hCoV-19/USA/MN-QDX-1020/2020\|EPI_ISL_571746\|2020-0 | ···················· | ···················· | ····················· | ···················· | ···················· | ···················· |
| L | 48 | hCoV-19/USA/MN-QDX-1022/2020\|EPI_ISL_571748\|2020-0 | ···················· | ···················· | ····················· | ···················· | ···················· | ···················· |
| L | 49 | hCoV-19/USA/MN-QDX-1023/2020\|EPI_ISL_571749\|2020-0 | ···················· | ···················· | ····················· | ···················· | ···················· | ···················· |
| L | 50 | hCoV-19/USA/MN-QDX-1024/2020\|EPI_ISL_571750\|2020-0 | ···················· | ···················· | ····················· | ···················· | ···················· | ···················· |
| L | 51 | hCoV-19/USA/MN-QDX-1025/2020\|EPI_ISL_571751\|2020-0 | ···················· | ···················· | ····················· | ···················· | ···················· | ···················· |
| L | 52 | hCoV-19/USA/MN-QDX-1026/2020\|EPI_ISL_571752\|2020-0 | ···················· | ···················· | ····················· | ···················· | ···················· | ···················· |
| L | 53 | hCoV-19/USA/MN-QDX-952/2020\|EPI_ISL_571753\|2020-03 | ···················· | ···················· | ····················· | ···················· | ···················· | ···················· |
| L | 54 | hCoV-19/USA/MN-QDX-953/2020\|EPI_ISL_571754\|2020-03 | ···················· | ···················· | ····················· | ···················· | ···················· | ···················· |
| L | 55 | hCoV-19/USA/MN-QDX-954/2020\|EPI_ISL_571755\|2020-03 | ···················· | ···················· | ····················· | ···················· | ···················· | ···················· |
| L | 56 | hCoV-19/USA/MN-QDX-955/2020\|EPI_ISL_571756\|2020-03 | ···················· | ···················· | ····················· | ···················· | ···················· | ···················· |
| L | 57 | hCoV-19/USA/MN-QDX-956/2020\|EPI_ISL_571757\|2020-03 | ···················· | ···················· | ····················· | ···················· | ···················· | ···················· |
| L | 58 | hCoV-19/USA/MN-QDX-957/2020\|EPI_ISL_571758\|2020-03 | ···················· | ···················· | ····················· | ···················· | ···················· | ···················· |
| L | 59 | hCoV-19/USA/MN-QDX-958/2020\|EPI_ISL_571759\|2020-03 | ···················· | ···················· | ····················· | ···················· | ···················· | ···················· |
| L | 60 | hCoV-19/USA/MN-QDX-959/2020\|EPI_ISL_571760\|2020-03 | ···················· | ···················· | ····················· | ···················· | ···················· | ···················· |
| L | 61 | hCoV-19/USA/MN-QDX-960/2020\|EPI_ISL_571761\|2020-03 | ···················· | ···················· | ····················· | ···················· | ···················· | ···················· |
| L | 62 | hCoV-19/USA/MN-QDX-961/2020\|EPI_ISL_571762\|2020-03 | ···················· | ···················· | ····················· | ···················· | ···················· | ···················· |
| L | 63 | hCoV-19/USA/MN-QDX-962/2020\|EPI_ISL_571763\|2020-03 | ···················· | ···················· | ····················· | ···················· | ···················· | ···················· |
| L | 64 | hCoV-19/USA/MN-QDX-965/2020\|EPI_ISL_571764\|2020-03 | ···················· | ···················· | ····················· | ···················· | ···················· | ···················· |
| L | 65 | hCoV-19/USA/MN-QDX-966/2020\|EPI_ISL_571765\|2020-03 | ···················· | ···················· | ····················· | ···················· | ···················· | ···················· |
| L | 66 | hCoV-19/USA/MN-QDX-967/2020\|EPI_ISL_571766\|2020-03 | ···················· | ···················· | ····················· | ···················· | ···················· | ···················· |
| L | 67 | hCoV-19/USA/MN-QDX-968/2020\|EPI_ISL_571767\|2020-03 | ···················· | ···················· | ····················· | ···················· | ···················· | ···················· |
| L | 68 | hCoV-19/USA/MN-QDX-969/2020\|EPI_ISL_571768\|2020-03 | ···················· | ···················· | ····················· | ···················· | ···················· | ···················· |
| L | 69 | hCoV-19/USA/MN-QDX-970/2020\|EPI_ISL_571769\|2020-03 | ···················· | ···················· | ····················· | ···················· | ···················· | ···················· |
| L | 70 | hCoV-19/USA/MN-QDX-971/2020\|EPI_ISL_571770\|2020-03 | ···················· | ···················· | ····················· | ···················· | ···················· | ···················· |
| L | 71 | hCoV-19/USA/MN-QDX-972/2020\|EPI_ISL_571771\|2020-03 | ···················· | ···················· | ····················· | ···················· | ···················· | ···················· |
| L | 72 | hCoV-19/USA/MN-QDX-973/2020\|EPI_ISL_571772\|2020-03 | ···················· | ···················· | ····················· | ···················· | ···················· | ···················· |
| L | 73 | hCoV-19/USA/MN-QDX-974/2020\|EPI_ISL_571773\|2020-03 | ···················· | ···················· | ····················· | ···················· | ···················· | ···················· |
| L | 74 | hCoV-19/USA/MN-QDX-975/2020\|EPI_ISL_571774\|2020-03 | ···················· | ···················· | ····················· | ···················· | ···················· | ···················· |
| L | 75 | hCoV-19/USA/MN-QDX-976/2020\|EPI_ISL_571775\|2020-03 | ···················· | ···················· | ····················· | ···················· | ···················· | ···················· |
| L | 76 | hCoV-19/USA/MN-QDX-977/2020\|EPI_ISL_571776\|2020-03 | ···················· | ···················· | ····················· | ···················· | ···················· | ···················· |
| L | 77 | hCoV-19/USA/MN-QDX-978/2020\|EPI_ISL_571777\|2020-03 | ···················· | ···················· | ····················· | ···················· | ···················· | ···················· |
| L | 78 | hCoV-19/USA/MN-QDX-979/2020\|EPI_ISL_571778\|2020-03 | ···················· | ···················· | ····················· | ···················· | ···················· | ···················· |
| L | 79 | hCoV-19/USA/MN-QDX-980/2020\|EPI_ISL_571779\|2020-03 | ···················· | ···················· | ····················· | ···················· | ···················· | ···················· |
| L | 80 | hCoV-19/USA/MN-QDX-981/2020\|EPI_ISL_571780\|2020-03 | ···················· | ···················· | ····················· | ···················· | ···················· | ···················· |
| L | 81 | hCoV-19/USA/MN-QDX-982/2020\|EPI_ISL_571781\|2020-03 | ···················· | ···················· | ····················· | ···················· | ···················· | ···················· |
| L | 82 | hCoV-19/USA/MN-QDX-983/2020\|EPI_ISL_571782\|2020-03 | ···················· | ···················· | ····················· | ···················· | ···················· | ···················· |
| L | 83 | hCoV-19/USA/MN-QDX-985/2020\|EPI_ISL_571783\|2020-03 | ···················· | ···················· | ····················· | ···················· | ···················· | ···················· |
| L | 84 | hCoV-19/USA/MN-QDX-986/2020\|EPI_ISL_571784\|2020-03 | ···················· | ···················· | ····················· | ···················· | ···················· | ···················· |
| L | 85 | hCoV-19/USA/MN-QDX-987/2020\|EPI_ISL_571785\|2020-03 | ···················· | ···················· | ····················· | ···················· | ···················· | ···················· |

```
L    86 hCoV-19/USA/MN-QDX-988/2020|EPI_ISL_571786|2020-03    ····················  ····················  ·····················  ····················  ····················  ····················
L    87 hCoV-19/USA/MN-QDX-989/2020|EPI_ISL_571787|2020-03    ····················  ····················  ·····················  ····················  ····················  ····················
L    88 hCoV-19/USA/MN-QDX-990/2020|EPI_ISL_571788|2020-03    ····················  ····················  ·····················  ····················  ····················  ····················
L    89 hCoV-19/USA/MN-QDX-991/2020|EPI_ISL_571789|2020-03    ····················  ····················  ·····················  ····················  ····················  ····················
L    90 hCoV-19/USA/MN-QDX-992/2020|EPI_ISL_571790|2020-03    ····················  ····················  ·····················  ····················  ····················  ····················
L    91 hCoV-19/USA/MN-QDX-994/2020|EPI_ISL_571791|2020-03    ····················  ····················  ·····················  ····················  ····················  ····················
L    92 hCoV-19/USA/MN-QDX-995/2020|EPI_ISL_571792|2020-03    ····················  ····················  ·····················  ····················  ····················  ····················
L    93 hCoV-19/USA/MN-QDX-996/2020|EPI_ISL_571793|2020-03    ····················  ····················  ·····················  ····················  ····················  ····················
L    94 hCoV-19/USA/MN-QDX-997/2020|EPI_ISL_571794|2020-03    ····················  ····················  ·····················  ····················  ····················  ····················
L    95 hCoV-19/USA/MN-QDX-998/2020|EPI_ISL_571795|2020-03    ····················  ····················  ·····················  ····················  ····················  ····················
L    96 hCoV-19/USA/MN-QDX-999/2020|EPI_ISL_571796|2020-03    ····················  ····················  ·····················  ····················  ····················  ····················
L    97 hCoV-19/USA/NV-QDX-1030/2020|EPI_ISL_571797|2020-0    ····················  ····················  ·····················  ····················  ····················  ····················
L    98 hCoV-19/USA/NV-QDX-1031/2020|EPI_ISL_571798|2020-0    ····················  ····················  ·····················  ····················  ····················  ····················
L    99 hCoV-19/USA/NY-QDX-950/2020|EPI_ISL_571799|2020-03    ····················  ····················  ·····················  ····················  ····················  ····················
L   100 hCoV-19/USA/NM-DOH-2020321214/2020|EPI_ISL_575290|    ····················  ····················  ·····················  ····················  ····················  ····················
L   101 hCoV-19/USA/NM-DOH-2020321219/2020|EPI_ISL_575291|    ····················  ····················  ·····················  ····················  ····················  ····················
L   102 hCoV-19/USA/NM-DOH-2020321223/2020|EPI_ISL_575293|    ····················  ····················  ·····················  ····················  ····················  ····················
L   103 hCoV-19/USA/NM-DOH-2020322589/2020|EPI_ISL_575295|    ····················  ····················  ·····················  ····················  ····················  ····················
L   104 hCoV-19/USA/NM-DOH-2020322592/2020|EPI_ISL_575296|    ····················  ····················  ·····················  ····················  ····················  ····················
L   105 hCoV-19/USA/NM-DOH-2020324092/2020|EPI_ISL_575297|    ····················  ····················  ·····················  ····················  ····················  ····················
L   106 hCoV-19/USA/NM-DOH-2020324349/2020|EPI_ISL_575298|    ····················  ····················  ·····················  ····················  ····················  ····················
L   107 hCoV-19/USA/NM-DOH-2020324378/2020|EPI_ISL_575299|    ····················  ····················  ·····················  ····················  ····················  ····················
L   108 hCoV-19/USA/NM-DOH-2020326789/2020|EPI_ISL_575300|    ····················  ····················  ·····················  ····················  ····················  ····················
L   109 hCoV-19/USA/NM-DOH-2020326823/2020|EPI_ISL_575302|    ····················  ····················  ·····················  ····················  ····················  ····················
L   110 hCoV-19/USA/NM-DOH-2020327022/2020|EPI_ISL_575303|    ····················  ·A··················  ·····················  ····················  ····················  ····················
L   111 hCoV-19/USA/NM-DOH-2020327026/2020|EPI_ISL_575305|    ····················  ·A··················  ·····················  ····················  ····················  ····················
L   112 hCoV-19/USA/NM-DOH-2020327064/2020|EPI_ISL_575306|    ····················  ····················  ·····················  ····················  ····················  ····················
L   113 hCoV-19/USA/NM-DOH-2020327107/2020|EPI_ISL_575307|    ····················  ····················  ·····················  ····················  ····················  ····················
L   114 hCoV-19/USA/NM-DOH-2020327163/2020|EPI_ISL_575309|    ····················  ····················  ·····················  ····················  ····················  ····················
L   115 hCoV-19/USA/NM-DOH-2020327257/2020|EPI_ISL_575310|    ····················  ····················  ·····················  ····················  ····················  ····················
L   116 hCoV-19/USA/NM-DOH-2020327510/2020|EPI_ISL_575311|    ····················  ····················  ·····················  ····················  ····················  ····················
L   117 hCoV-19/USA/NM-DOH-2020327831/2020|EPI_ISL_575312|    ····················  ····················  ·····················  ····················  ····················  ····················
L   118 hCoV-19/Beijing/NPRC0004/2020|EPI_ISL_575330|2020-    ····················  ····················  ·····················  ····················  ····················  ····················
L   119 hCoV-19/England/ALDP-9D783E/2020|EPI_ISL_576589|20    ····················  ····················  ·····················  ····················  ····················  ····················
L   120 hCoV-19/England/ALDP-9D784D/2020|EPI_ISL_576590|20    ····················  ····················  ·····················  ····················  ····················  ····················
L   121 hCoV-19/England/ALDP-9D785C/2020|EPI_ISL_576591|20    ····················  ····················  ·····················  ····················  ····················  ····················
L   122 hCoV-19/England/ALDP-9D787A/2020|EPI_ISL_576592|20    ····················  ····················  ·····················  ····················  ····················  ····················
L   123 hCoV-19/England/ALDP-9D7889/2020|EPI_ISL_576593|20    ····················  ····················  ·····················  ····················  ····················  ····················
L   124 hCoV-19/England/ALDP-9D78A7/2020|EPI_ISL_576594|20    ····················  ····················  ·····················  ····················  ····················  ····················
L   125 hCoV-19/England/ALDP-9D78B6/2020|EPI_ISL_576595|20    ····················  ····················  ·····················  ····················  ····················  ····················
L   126 hCoV-19/England/ALDP-9D78C5/2020|EPI_ISL_576596|20    ····················  ····················  ·····················  ····················  ····················  ····················
L   127 hCoV-19/England/ALDP-9D78D4/2020|EPI_ISL_576597|20    ····················  ····················  ·····················  ····················  ····················  ····················
L   128 hCoV-19/England/ALDP-9D790E/2020|EPI_ISL_576598|20    ····················  ····················  ·····················  ····················  ····················  ····················
L   129 hCoV-19/England/ALDP-9D792C/2020|EPI_ISL_576599|20    ····················  ·A··················  ·····················  ····················  ····················  ··A·················
L   130 hCoV-19/England/ALDP-9D7CBA/2020|EPI_ISL_576622|20    ····················  ····················  ·····················  ····················  ····················  ····················
L   131 hCoV-19/England/ALDP-9D7CC9/2020|EPI_ISL_576623|20    ····················  ····················  ·····················  ····················  ····················  ····················
L   132 hCoV-19/England/ALDP-9D7CE7/2020|EPI_ISL_576624|20    ····················  ····················  ·····················  ····················  ····················  ····················
L   133 hCoV-19/England/ALDP-9D7D20/2020|EPI_ISL_576626|20    ····················  ····················  ·····················  ····················  ····················  ····················
L   134 hCoV-19/England/ALDP-9D7D7B/2020|EPI_ISL_576628|20    ····················  ····················  ·····················  ····················  ····················  ····················
L   135 hCoV-19/England/ALDP-9D7DD5/2020|EPI_ISL_576630|20    ····················  ····················  ·····················  ····················  ····················  ····················
L   136 hCoV-19/England/ALDP-9D7DE4/2020|EPI_ISL_576631|20    ····················  ····················  ·····················  ····················  ····················  ····················
L   137 hCoV-19/England/ALDP-9D7DF3/2020|EPI_ISL_576632|20    ····················  ····················  ·····················  ····················  ····················  ····················
L   138 hCoV-19/England/ALDP-9D7E1E/2020|EPI_ISL_576633|20    ····················  ····················  ·····················  ····················  ····················  ····················
L   139 hCoV-19/England/ALDP-9D7E2D/2020|EPI_ISL_576634|20    ····················  ····················  ·····················  ····················  ····················  ····················
L   140 hCoV-19/England/ALDP-9D7E3C/2020|EPI_ISL_576635|20    ····················  ····················  ·····················  ····················  ····················  ····················
L   141 hCoV-19/England/ALDP-9D7FA2/2020|EPI_ISL_576636|20    ····················  ····················  ·····················  ····················  ····················  ····················
L   142 hCoV-19/England/ALDP-9D7FC0/2020|EPI_ISL_576637|20    ····················  ·A··················  ·····················  ····················  ····················  ··A·················
L   143 hCoV-19/England/ALDP-9D7FDF/2020|EPI_ISL_576638|20    ····················  ·A··················  ·····················  ····················  ····················  ··A·················
L   144 hCoV-19/England/ALDP-9D80F9/2020|EPI_ISL_576639|20    ····················  ····················  ·····················  ····················  ····················  ····················
L   145 hCoV-19/England/ALDP-9D8150/2020|EPI_ISL_576640|20    ····················  ····················  ·····················  ····················  ····················  ····················
L   146 hCoV-19/England/ALDP-9D824E/2020|EPI_ISL_576641|20    ····················  ·A··················  ·····················  ····················  ····················  ··A·················
L   147 hCoV-19/England/ALDP-9D826C/2020|EPI_ISL_576642|20    ····················  ····················  ·····················  ····················  ····················  ····················
L   148 hCoV-19/England/ALDP-9D828A/2020|EPI_ISL_576643|20    ····················  ····················  ·····················  ····················  ····················  ····················
L   149 hCoV-19/England/ALDP-9D8CC8/2020|EPI_ISL_576661|20    ····················  ····················  ·····················  ····················  ····················  ····················
L   150 hCoV-19/Netherlands/ZH-EMC-525/2020|EPI_ISL_577953    ····················  ····················  ·····················  ····················  ····················  ····················
L   151 hCoV-19/Netherlands/ZH-EMC-526/2020|EPI_ISL_577954    ····················  ····················  ·····················  ····················  ····················  ····················
L   152 hCoV-19/Netherlands/ZH-EMC-527/2020|EPI_ISL_577955    ····················  ····················  ·····················  ····················  ····················  ····················
L   153 hCoV-19/Netherlands/ZH-EMC-528/2020|EPI_ISL_577956    ····················  ····················  ·····················  ····················  ····················  ····················
L   154 hCoV-19/Netherlands/ZH-EMC-529/2020|EPI_ISL_577957    ····················  ····················  ·····················  ····················  ····················  ····················
L   155 hCoV-19/Netherlands/ZH-EMC-530/2020|EPI_ISL_577958    ····················  ····················  ·····················  ····················  ····················  ····················
L   156 hCoV-19/Netherlands/ZH-EMC-531/2020|EPI_ISL_577959    ····················  ····················  ·····················  ····················  ····················  ····················
L   157 hCoV-19/Netherlands/ZH-EMC-532/2020|EPI_ISL_577960    ····················  ····················  ·····················  ····················  ····················  ····················
L   158 hCoV-19/Netherlands/ZH-EMC-533/2020|EPI_ISL_577961    ····················  ····················  ·····················  ····················  ····················  ····················
L   159 hCoV-19/Netherlands/ZH-EMC-535/2020|EPI_ISL_577963    ····················  ····················  ·····················  ····················  ····················  ····················
L   160 hCoV-19/Netherlands/ZH-EMC-536/2020|EPI_ISL_577964    ····················  ····················  ·····················  ····················  ····················  ····················
L   161 hCoV-19/Netherlands/ZH-EMC-538/2020|EPI_ISL_577966    ····················  ····················  ·····················  ····················  ····················  ····················
L   162 hCoV-19/Netherlands/ZH-EMC-539/2020|EPI_ISL_577967    ····················  ····················  ·····················  ····················  ······T·············  ····················
L   163 hCoV-19/Netherlands/ZH-EMC-541/2020|EPI_ISL_577969    ····················  ····················  ·····················  ····················  ····················  ····················
L   164 hCoV-19/Netherlands/ZH-EMC-542/2020|EPI_ISL_577970    ····················  ····················  ·····················  ····················  ····················  ····················
L   165 hCoV-19/Netherlands/ZH-EMC-543/2020|EPI_ISL_577971    ····················  ····················  ·····················  ····················  ····················  ····················
```

| | | | | | | | | |
|---|---|---|---|---|---|---|---|---|
| L | 166 | hCoV-19/Netherlands/ZH-EMC-544/2020\|EPI_ISL_577972 | ···················· | ···················· | ····················· | ···················· | ···················· | ···················· |
| L | 167 | hCoV-19/Netherlands/ZH-EMC-545/2020\|EPI_ISL_577973 | ···················· | ···················· | ····················· | ···················· | ···················· | ···················· |
| L | 168 | hCoV-19/England/QEUH-9DD2B2/2020\|EPI_ISL_581100\|20 | ···················· | ···················· | ····················· | ···················· | ···················· | ···················· |
| L | 169 | hCoV-19/England/QEUH-9DD319/2020\|EPI_ISL_581101\|20 | ···················· | ···················· | ····················· | ···················· | ···················· | ···················· |
| L | 170 | hCoV-19/England/QEUH-9DC848/2020\|EPI_ISL_581102\|20 | ···················· | ·A·················· | ····················· | ···················· | ···················· | ···A················ |
| L | 171 | hCoV-19/England/MILK-9E135E/2020\|EPI_ISL_581103\|20 | ···················· | ·A·················· | ····················· | ···················· | ···················· | ···A················ |
| L | 172 | hCoV-19/England/QEUH-9DAE1B/2020\|EPI_ISL_581104\|20 | ···················· | ···················· | ····················· | ···················· | ···················· | ···················· |
| L | 173 | hCoV-19/England/QEUH-9DC8EE/2020\|EPI_ISL_581105\|20 | ···················· | ···················· | ····················· | ···················· | ···················· | ···················· |
| L | 174 | hCoV-19/Wales/MILK-9E1039/2020\|EPI_ISL_581106\|2020 | ···················· | ···················· | ····················· | ···················· | ···················· | ···················· |
| L | 175 | hCoV-19/England/MILK-9E0EA9/2020\|EPI_ISL_581107\|20 | ···················· | ···················· | ····················· | ···················· | ···················· | ···················· |
| L | 176 | hCoV-19/England/QEUH-9DC796/2020\|EPI_ISL_581108\|20 | ···················· | ···················· | ····················· | ···················· | ···················· | ···················· |
| L | 177 | hCoV-19/England/QEUH-9DD2EF/2020\|EPI_ISL_581109\|20 | ···················· | ···················· | ····················· | ···················· | ···················· | ···················· |
| L | 178 | hCoV-19/England/QEUH-9DAE2A/2020\|EPI_ISL_581110\|20 | ···················· | ···················· | ····················· | ···················· | ···················· | ···················· |
| L | 179 | hCoV-19/England/MILK-9E1242/2020\|EPI_ISL_581111\|20 | ···················· | ···················· | ····················· | ···················· | ···················· | ···················· |
| L | 180 | hCoV-19/England/QEUH-9DCA24/2020\|EPI_ISL_581112\|20 | ···················· | ···················· | ····················· | ···················· | ···················· | ···················· |
| L | 181 | hCoV-19/England/MILK-9E237B/2020\|EPI_ISL_581113\|20 | ···················· | ···················· | ····················· | ···················· | ···················· | ···················· |
| L | 182 | hCoV-19/England/MILK-9E21F9/2020\|EPI_ISL_581114\|20 | ···················· | ···················· | ····················· | ···················· | ···················· | ···················· |
| L | 183 | hCoV-19/England/MILK-9E1DC9/2020\|EPI_ISL_581115\|20 | ···················· | ···················· | ····················· | ···················· | ······T············· | ···················· |
| L | 184 | hCoV-19/England/MILK-9E2EC5/2020\|EPI_ISL_581116\|20 | ···················· | ···················· | ····················· | ···················· | ···················· | ···················· |
| L | 185 | hCoV-19/England/MILK-9E2C25/2020\|EPI_ISL_581118\|20 | ···················· | ·A·················· | ····················· | ···················· | ···················· | ···A················ |
| L | 186 | hCoV-19/England/MILK-9E1D05/2020\|EPI_ISL_581119\|20 | ···················· | ···················· | ····················· | ···················· | ···················· | ···················· |
| L | 187 | hCoV-19/Wales/MILK-9E2065/2020\|EPI_ISL_581120\|2020 | ···················· | ···················· | ····················· | ···················· | ···················· | ···················· |
| L | 188 | hCoV-19/England/MILK-9E1D41/2020\|EPI_ISL_581121\|20 | ···················· | ···················· | ····················· | ···················· | ···················· | ···················· |
| L | 189 | hCoV-19/Wales/MILK-9E2180/2020\|EPI_ISL_581122\|2020 | ···················· | ···················· | ····················· | ···················· | ···················· | ···················· |
| L | 190 | hCoV-19/England/MILK-9E2223/2020\|EPI_ISL_581123\|20 | ···················· | ···················· | ····················· | ···················· | ···················· | ···················· |
| L | 191 | hCoV-19/England/MILK-9E336B/2020\|EPI_ISL_581124\|20 | ···················· | ···················· | ····················· | ···················· | ···················· | ···················· |
| L | 192 | hCoV-19/Wales/MILK-9E1E5D/2020\|EPI_ISL_581126\|2020 | ···················· | ···················· | ····················· | ···················· | ···················· | ···················· |
| L | 193 | hCoV-19/England/QEUH-9DEF14/2020\|EPI_ISL_581127\|20 | ···················· | ·A·················· | ····················· | ···················· | ···················· | ···A················ |
| L | 194 | hCoV-19/Wales/MILK-9E2D04/2020\|EPI_ISL_581128\|2020 | ···················· | ···················· | ····················· | ···················· | ···················· | ···················· |
| L | 195 | hCoV-19/England/MILK-9E2A58/2020\|EPI_ISL_581129\|20 | ···················· | ···················· | ····················· | ···················· | ···················· | ···················· |
| L | 196 | hCoV-19/England/MILK-9E31BC/2020\|EPI_ISL_581131\|20 | ···················· | ···················· | ····················· | ···················· | ···················· | ···················· |
| L | 197 | hCoV-19/Canada/MB-NML-1039/2020\|EPI_ISL_582400\|202 | ···················· | ···················· | ····················· | ···················· | ···················· | ···················· |
| L | 198 | hCoV-19/Canada/MB-NML-1041/2020\|EPI_ISL_582401\|202 | ···················· | ···················· | ····················· | ···················· | ···················· | ···················· |
| L | 199 | hCoV-19/Canada/MB-NML-1042/2020\|EPI_ISL_582402\|202 | ···················· | ···················· | ····················· | ···················· | ···················· | ···················· |
| L | 200 | hCoV-19/Canada/MB-NML-1043/2020\|EPI_ISL_582403\|202 | ···················· | ···················· | ····················· | ···················· | ···················· | ···················· |
| L | 201 | hCoV-19/Canada/MB-NML-1044/2020\|EPI_ISL_582404\|202 | ···················· | ···················· | ····················· | ···················· | ···················· | ···················· |
| L | 202 | hCoV-19/Canada/MB-NML-1045/2020\|EPI_ISL_582405\|202 | ···················· | ·A·················· | ····················· | ···················· | ···················· | ···A················ |
| L | 203 | hCoV-19/Canada/MB-NML-1046/2020\|EPI_ISL_582406\|202 | ···················· | ···················· | ····················· | ···················· | ···················· | ···················· |
| L | 204 | hCoV-19/Canada/MB-NML-1048/2020\|EPI_ISL_582407\|202 | ···················· | ···················· | ····················· | ···················· | ···················· | ···················· |
| L | 205 | hCoV-19/Canada/MB-NML-1049/2020\|EPI_ISL_582408\|202 | ···················· | ···················· | ····················· | ···················· | ···················· | ···················· |
| L | 206 | hCoV-19/Canada/MB-NML-1050/2020\|EPI_ISL_582409\|202 | ···················· | ···················· | ····················· | ···················· | ···················· | ···················· |
| L | 207 | hCoV-19/Canada/MB-NML-1051/2020\|EPI_ISL_582410\|202 | ···················· | ···················· | ····················· | ···················· | ···················· | ···················· |
| L | 208 | hCoV-19/Canada/MB-NML-1052/2020\|EPI_ISL_582411\|202 | ···················· | ···················· | ····················· | ···················· | ···················· | ···················· |
| L | 209 | hCoV-19/Canada/MB-NML-1053/2020\|EPI_ISL_582412\|202 | ···················· | ·A·················· | ····················· | ···················· | ···················· | ···A················ |
| L | 210 | hCoV-19/Canada/MB-NML-1054/2020\|EPI_ISL_582413\|202 | ···················· | ···················· | ····················· | ···················· | ···················· | ···················· |
| L | 211 | hCoV-19/Canada/MB-NML-1055/2020\|EPI_ISL_582414\|202 | ···················· | ···················· | ····················· | ···················· | ···················· | ···················· |
| L | 212 | hCoV-19/Canada/MB-NML-1060/2020\|EPI_ISL_582416\|202 | ···················· | ·A·················· | ····················· | ···················· | ···················· | ···A················ |
| L | 213 | hCoV-19/Canada/MB-NML-1061/2020\|EPI_ISL_582417\|202 | ···················· | ·A·················· | ····················· | ···················· | ···················· | ···A················ |
| L | 214 | hCoV-19/Canada/MB-NML-1063/2020\|EPI_ISL_582418\|202 | ···················· | ···················· | ····················· | ···················· | ···················· | ···················· |
| L | 215 | hCoV-19/Canada/MB-NML-1064/2020\|EPI_ISL_582419\|202 | ···················· | ···················· | ····················· | ···················· | ···················· | ···················· |
| L | 216 | hCoV-19/Canada/MB-NML-1065/2020\|EPI_ISL_582420\|202 | ···················· | ···················· | ····················· | ···················· | ···················· | ···················· |
| L | 217 | hCoV-19/Canada/MB-NML-1067/2020\|EPI_ISL_582422\|202 | ···················· | ···················· | ····················· | ···················· | ···················· | ···················· |
| L | 218 | hCoV-19/Canada/MB-NML-1069/2020\|EPI_ISL_582423\|202 | ···················· | ···················· | ····················· | ···················· | ···················· | ···················· |
| L | 219 | hCoV-19/Canada/MB-NML-1070/2020\|EPI_ISL_582424\|202 | ···················· | ·A·················· | ····················· | ···················· | ···················· | ···A················ |
| L | 220 | hCoV-19/Canada/MB-NML-1071/2020\|EPI_ISL_582425\|202 | ···················· | ···················· | ····················· | ···················· | ···················· | ···················· |
| L | 221 | hCoV-19/Canada/MB-NML-1073/2020\|EPI_ISL_582426\|202 | ···················· | ···················· | ····················· | ···················· | ···················· | ···················· |
| L | 222 | hCoV-19/Canada/MB-NML-1075/2020\|EPI_ISL_582427\|202 | ···················· | ···················· | ····················· | ···················· | ···················· | ···················· |
| L | 223 | hCoV-19/Canada/MB-NML-1076/2020\|EPI_ISL_582428\|202 | ···················· | ·A·················· | ····················· | ···················· | ···················· | ···A················ |
| L | 224 | hCoV-19/Canada/MB-NML-1077/2020\|EPI_ISL_582429\|202 | ···················· | ·A·················· | ····················· | ···················· | ···················· | ···A················ |
| L | 225 | hCoV-19/Canada/MB-NML-1078/2020\|EPI_ISL_582430\|202 | ···················· | ·A·················· | ····················· | ···················· | ···················· | ···A················ |
| L | 226 | hCoV-19/Canada/MB-NML-1079/2020\|EPI_ISL_582431\|202 | ···················· | ···················· | ····················· | ···················· | ···················· | ···················· |
| L | 227 | hCoV-19/Canada/MB-NML-1080/2020\|EPI_ISL_582432\|202 | ···················· | ·A·················· | ····················· | ···················· | ···················· | ···A················ |
| L | 228 | hCoV-19/Canada/MB-NML-1082/2020\|EPI_ISL_582433\|202 | ···················· | ···················· | ····················· | ···················· | ···················· | ···················· |
| L | 229 | hCoV-19/Canada/MB-NML-1083/2020\|EPI_ISL_582434\|202 | ···················· | ·A·················· | ····················· | ···················· | ···················· | ···A················ |
| L | 230 | hCoV-19/Canada/MB-NML-1085/2020\|EPI_ISL_582435\|202 | ···················· | ·A·················· | ····················· | ···················· | ···················· | ···A················ |
| L | 231 | hCoV-19/Canada/MB-NML-1086/2020\|EPI_ISL_582436\|202 | ···················· | ···················· | ····················· | ···················· | ···················· | ···················· |
| L | 232 | hCoV-19/Canada/MB-NML-1087/2020\|EPI_ISL_582437\|202 | ···················· | ···················· | ····················· | ···················· | ···················· | ···················· |
| L | 233 | hCoV-19/Canada/MB-NML-1089/2020\|EPI_ISL_582438\|202 | ···················· | ···················· | ····················· | ···················· | ···················· | ···················· |
| L | 234 | hCoV-19/Canada/MB-NML-1090/2020\|EPI_ISL_582439\|202 | ···················· | ···················· | ····················· | ···················· | ···················· | ···················· |
| L | 235 | hCoV-19/Canada/MB-NML-1091/2020\|EPI_ISL_582440\|202 | ···················· | ·N·················· | ····················· | ···················· | ···················· | ···N················ |
| L | 236 | hCoV-19/Canada/MB-NML-1092/2020\|EPI_ISL_582441\|202 | ···················· | ·A·················· | ····················· | ···················· | ···················· | ···A················ |
| L | 237 | hCoV-19/Canada/MB-NML-1093/2020\|EPI_ISL_582442\|202 | ···················· | ···················· | ····················· | ···················· | ···················· | ···················· |
| L | 238 | hCoV-19/Canada/MB-NML-1096/2020\|EPI_ISL_582443\|202 | ···················· | ·A·················· | ····················· | ···················· | ···················· | ···A················ |
| L | 239 | hCoV-19/Canada/MB-NML-1100/2020\|EPI_ISL_582445\|202 | ···················· | ·A·················· | ····················· | ···················· | ···················· | ···A················ |
| L | 240 | hCoV-19/Canada/MB-NML-1101/2020\|EPI_ISL_582446\|202 | ···················· | ·A·················· | ····················· | ···················· | ···················· | ···A················ |
| L | 241 | hCoV-19/Canada/MB-NML-1102/2020\|EPI_ISL_582447\|202 | ···················· | ·A·················· | ····················· | ···················· | ···················· | ···A················ |
| L | 242 | hCoV-19/Canada/MB-NML-1104/2020\|EPI_ISL_582448\|202 | ···················· | ·A·················· | ····················· | ···················· | ···················· | ···A················ |
| L | 243 | hCoV-19/Canada/MB-NML-1107/2020\|EPI_ISL_582449\|202 | ···················· | ···················· | ····················· | ···················· | ···················· | ···················· |
| L | 244 | hCoV-19/Canada/MB-NML-1110/2020\|EPI_ISL_582450\|202 | ···················· | ·A·················· | ····················· | ···················· | ···················· | ···A················ |
| L | 245 | hCoV-19/Canada/MB-NML-1111/2020\|EPI_ISL_582451\|202 | ···················· | ···················· | ····················· | ···················· | ···················· | ···················· |

| | | | | | | | | |
|---|---|---|---|---|---|---|---|---|
| L | 246 | hCoV-19/Canada/MB-NML-1112/2020\|EPI_ISL_582452\|202 | ···················· | ·A·················· | ···················· | ···················· | ···················· | ···A················ |
| L | 247 | hCoV-19/Canada/MB-NML-1116/2020\|EPI_ISL_582453\|202 | ···················· | ·A·················· | ···················· | ···················· | ···················· | ···A················ |
| L | 248 | hCoV-19/Canada/MB-NML-1117/2020\|EPI_ISL_582454\|202 | ···················· | ···················· | ···················· | ···················· | ···················· | ···················· |
| L | 249 | hCoV-19/Canada/MB-NML-1119/2020\|EPI_ISL_582455\|202 | ···················· | ·A·················· | ···················· | ···················· | ···················· | ···A················ |
| L | 250 | hCoV-19/Canada/MB-NML-1121/2020\|EPI_ISL_582456\|202 | ···················· | ···················· | ···················· | ···················· | ···················· | ···················· |
| L | 251 | hCoV-19/Canada/MB-NML-1122/2020\|EPI_ISL_582457\|202 | ···················· | ·A·················· | ···················· | ···················· | ···················· | ···A················ |
| L | 252 | hCoV-19/Canada/MB-NML-1124/2020\|EPI_ISL_582458\|202 | ···················· | ·A·················· | ···················· | ···················· | ···················· | ···A················ |
| L | 253 | hCoV-19/Canada/MB-NML-1125/2020\|EPI_ISL_582459\|202 | ···················· | ·A·················· | ···················· | ···················· | ···················· | ···A················ |
| L | 254 | hCoV-19/Canada/MB-NML-1128/2020\|EPI_ISL_582460\|202 | ···················· | ·A·················· | ···················· | ···················· | ···················· | ···A················ |
| L | 255 | hCoV-19/Canada/MB-NML-1130/2020\|EPI_ISL_582461\|202 | ···················· | ·A·················· | ···················· | ···················· | ···················· | ···A················ |
| L | 256 | hCoV-19/Canada/MB-NML-1131/2020\|EPI_ISL_582462\|202 | ···················· | ···················· | ···················· | ···················· | ···················· | ···················· |
| L | 257 | hCoV-19/Wales/PHWC-16BFE7/2020\|EPI_ISL_574000\|2020 | ···················· | ···················· | ···················· | ···················· | ···················· | ···················· |
| L | 258 | hCoV-19/Wales/PHWC-16BFF6/2020\|EPI_ISL_574001\|2020 | ···················· | ···················· | ···················· | ···················· | ···················· | ···················· |
| L | 260 | hCoV-19/Wales/PHWC-16C010/2020\|EPI_ISL_574003\|2020 | ···················· | ···················· | ···················· | ···················· | ···················· | ···················· |
| L | 261 | hCoV-19/Wales/PHWC-16C02F/2020\|EPI_ISL_574004\|2020 | ···················· | ···················· | ···················· | ···················· | ···················· | ···················· |
| L | 262 | hCoV-19/Wales/PHWC-16C04D/2020\|EPI_ISL_574005\|2020 | ···················· | ···················· | ···················· | ···················· | ······T············· | ···················· |
| L | 264 | hCoV-19/Wales/PHWC-16C06B/2020\|EPI_ISL_574007\|2020 | ···················· | ···················· | ···················· | ···················· | ···················· | ···················· |
| L | 265 | hCoV-19/Wales/PHWC-16C098/2020\|EPI_ISL_574008\|2020 | ···················· | ···················· | ···················· | ···················· | ···················· | ···················· |
| L | 266 | hCoV-19/Wales/PHWC-16C0B6/2020\|EPI_ISL_574009\|2020 | ···················· | ···················· | ···················· | ···················· | ···················· | ···················· |
| L | 267 | hCoV-19/Wales/PHWC-16C0E3/2020\|EPI_ISL_574010\|2020 | ···················· | ···················· | ···················· | ···················· | ···················· | ···················· |
| L | 268 | hCoV-19/Wales/PHWC-16C0F2/2020\|EPI_ISL_574011\|2020 | ···················· | ···················· | ···················· | ···················· | ···················· | ···················· |
| L | 269 | hCoV-19/Wales/PHWC-16C11D/2020\|EPI_ISL_574012\|2020 | ···················· | ···················· | ···················· | ···················· | ···················· | ···················· |
| L | 270 | hCoV-19/Wales/PHWC-16C12C/2020\|EPI_ISL_574013\|2020 | ···················· | ···················· | ···················· | ···················· | ···················· | ···················· |
| L | 271 | hCoV-19/Wales/PHWC-16C13B/2020\|EPI_ISL_574014\|2020 | ···················· | ···················· | ···················· | ···················· | ···················· | ···················· |
| L | 272 | hCoV-19/Wales/PHWC-16C14A/2020\|EPI_ISL_574015\|2020 | ···················· | ···················· | ···················· | ···················· | ···················· | ···················· |
| L | 273 | hCoV-19/Wales/PHWC-16C159/2020\|EPI_ISL_574016\|2020 | ···················· | ···················· | ···················· | ···················· | ···················· | ···················· |
| L | 274 | hCoV-19/Wales/PHWC-16C168/2020\|EPI_ISL_574017\|2020 | ···················· | ···················· | ···················· | ···················· | ···················· | ···················· |
| L | 275 | hCoV-19/Wales/PHWC-16C186/2020\|EPI_ISL_574018\|2020 | ···················· | ···················· | ···················· | ···················· | ···················· | ···················· |
| L | 276 | hCoV-19/Wales/PHWC-16C195/2020\|EPI_ISL_574019\|2020 | ···················· | ···················· | ···················· | ···················· | ···················· | ···················· |
| L | 277 | hCoV-19/Wales/PHWC-16C1A4/2020\|EPI_ISL_574020\|2020 | ···················· | ···················· | ···················· | ···················· | ···················· | ···················· |
| L | 278 | hCoV-19/Wales/PHWC-16C1B3/2020\|EPI_ISL_574021\|2020 | ···················· | ···················· | ···················· | ···················· | ···················· | ···················· |
| L | 279 | hCoV-19/Wales/PHWC-16C1C2/2020\|EPI_ISL_574022\|2020 | ···················· | ···················· | ···················· | ···················· | ···················· | ···················· |
| L | 281 | hCoV-19/Wales/PHWC-16C1FF/2020\|EPI_ISL_574024\|2020 | ···················· | ···················· | ···················· | ···················· | N··················· | ···················· |
| L | 282 | hCoV-19/Wales/PHWC-16C20B/2020\|EPI_ISL_574025\|2020 | ···················· | ···················· | ···················· | ···················· | ···················· | ···················· |
| L | 283 | hCoV-19/Wales/PHWC-16C21A/2020\|EPI_ISL_574026\|2020 | ···················· | ···················· | ···················· | ···················· | ···················· | ···················· |
| L | 284 | hCoV-19/Wales/PHWC-16C229/2020\|EPI_ISL_574027\|2020 | ···················· | ···················· | ···················· | ···················· | ···················· | ···················· |
| L | 285 | hCoV-19/Wales/PHWC-16C238/2020\|EPI_ISL_574028\|2020 | ···················· | ···················· | ···················· | ···················· | ···················· | ···················· |
| L | 286 | hCoV-19/Wales/PHWC-16C256/2020\|EPI_ISL_574029\|2020 | ···················· | ···················· | ···················· | ···················· | ···················· | ···················· |
| L | 287 | hCoV-19/Wales/PHWC-16C274/2020\|EPI_ISL_574030\|2020 | ···················· | ···················· | ···················· | ···················· | ···················· | ···················· |
| L | 288 | hCoV-19/Wales/PHWC-16C283/2020\|EPI_ISL_574031\|2020 | ···················· | ···················· | ···················· | ···················· | ···················· | ···················· |
| L | 290 | hCoV-19/Wales/PHWC-16C2A1/2020\|EPI_ISL_574033\|2020 | ···················· | ···················· | ···················· | ···················· | ···················· | ···················· |
| L | 291 | hCoV-19/Wales/PHWC-16C2B0/2020\|EPI_ISL_574034\|2020 | ···················· | ···················· | ···················· | ···················· | ···················· | ···················· |
| L | 293 | hCoV-19/Wales/PHWC-16C2DE/2020\|EPI_ISL_574036\|2020 | ···················· | ···················· | ···················· | ···················· | ···················· | ···················· |
| L | 294 | hCoV-19/Wales/PHWC-16C2ED/2020\|EPI_ISL_574037\|2020 | ···················· | ···················· | ···················· | ···················· | ···················· | ···················· |
| L | 295 | hCoV-19/Wales/PHWC-16C2FC/2020\|EPI_ISL_574038\|2020 | ···················· | ···················· | ···················· | ···················· | ···················· | ···················· |
| L | 297 | hCoV-19/Wales/PHWC-16C317/2020\|EPI_ISL_574040\|2020 | ···················· | ···················· | ···················· | ···················· | ···················· | ···················· |
| L | 298 | hCoV-19/Wales/PHWC-16C326/2020\|EPI_ISL_574041\|2020 | ···················· | ···················· | ···················· | ···················· | ···················· | ···················· |
| L | 299 | hCoV-19/Wales/PHWC-16C335/2020\|EPI_ISL_574042\|2020 | ···················· | ···················· | ···················· | ···················· | ···················· | ···················· |
| L | 300 | hCoV-19/Wales/PHWC-16C344/2020\|EPI_ISL_574043\|2020 | ···················· | ···················· | ···················· | ···················· | ···················· | ···················· |
| L | 301 | hCoV-19/Wales/PHWC-16C353/2020\|EPI_ISL_574044\|2020 | ···················· | ···················· | ···················· | ···················· | ···················· | ···················· |
| L | 302 | hCoV-19/Wales/PHWC-16C362/2020\|EPI_ISL_574045\|2020 | ···················· | ···················· | ···················· | ···················· | ···················· | ···················· |
| L | 303 | hCoV-19/Wales/PHWC-16C371/2020\|EPI_ISL_574046\|2020 | ···················· | ···················· | ···················· | ···················· | ···················· | ···················· |
| L | 304 | hCoV-19/Wales/PHWC-16C380/2020\|EPI_ISL_574047\|2020 | ···················· | ···················· | ···················· | ···················· | ···················· | ···················· |
| L | 305 | hCoV-19/USA/NM-DOH-2020327863/2020\|EPI_ISL_575313\| | ···················· | ···················· | ···················· | ···················· | ···················· | ···················· |
| L | 306 | hCoV-19/USA/NM-DOH-2020328229/2020\|EPI_ISL_575314\| | ···················· | ···················· | ···················· | ···················· | ···················· | ···················· |
| L | 307 | hCoV-19/USA/NM-DOH-2020328244/2020\|EPI_ISL_575315\| | ···················· | ···················· | ···················· | ···················· | ···················· | ···················· |
| L | 308 | hCoV-19/USA/NM-DOH-2020328404/2020\|EPI_ISL_575316\| | ···················· | ···················· | ···················· | ···················· | ···················· | ···················· |
| L | 309 | hCoV-19/USA/NM-DOH-2020328407/2020\|EPI_ISL_575317\| | ···················· | ···················· | ···················· | ···················· | ···················· | ···················· |
| L | 310 | hCoV-19/USA/NM-DOH-2020328416/2020\|EPI_ISL_575318\| | ···················· | ···················· | ···················· | ···················· | ···················· | ···················· |
| L | 311 | hCoV-19/USA/NM-DOH-2020328523/2020\|EPI_ISL_575319\| | ···················· | ···················· | ···················· | ···················· | ···················· | ···················· |
| L | 312 | hCoV-19/USA/NM-DOH-2020328525/2020\|EPI_ISL_575320\| | ···················· | ···················· | ···················· | ···················· | ···················· | ···················· |
| L | 313 | hCoV-19/USA/NM-DOH-2020328640/2020\|EPI_ISL_575321\| | ···················· | ···················· | ···················· | ···················· | ···················· | ···················· |
| L | 314 | hCoV-19/USA/NM-DOH-2020328645/2020\|EPI_ISL_575322\| | ···················· | ···················· | ···················· | ···················· | ···················· | ···················· |
| L | 315 | hCoV-19/USA/NM-DOH-2020328767/2020\|EPI_ISL_575323\| | ···················· | ·A·················· | ···················· | ···················· | ···················· | ···················· |
| L | 316 | hCoV-19/USA/NM-DOH-2020329070/2020\|EPI_ISL_575324\| | ···················· | ···················· | ···················· | ···················· | ···················· | ···················· |
| L | 317 | hCoV-19/USA/NM-DOH-2020329281/2020\|EPI_ISL_575325\| | ···················· | ···················· | ···················· | ···················· | ···················· | ···················· |
| L | 318 | hCoV-19/USA/NM-DOH-2020329356/2020\|EPI_ISL_575326\| | ···················· | ·A·················· | ···················· | ···················· | ···················· | ···················· |
| L | 319 | hCoV-19/Indonesia/YO-UGM-00061/2020\|EPI_ISL_575331 | ···················· | ···················· | ···················· | ···················· | ···················· | ···················· |
| L | 320 | hCoV-19/Israel/CVL-s2049/2020\|EPI_ISL_575332\|2020- | ···················· | ·A·················· | ···················· | ···················· | ···················· | ···················· |
| L | 321 | hCoV-19/Israel/CVL-s2057/2020\|EPI_ISL_575333\|2020- | ···················· | ·A·················· | ···················· | ···················· | ···················· | ···················· |
| L | 322 | hCoV-19/Israel/CVL-s2064/2020\|EPI_ISL_575334\|2020- | ···················· | ·A·················· | ···················· | ···················· | ···················· | ···················· |
| L | 323 | hCoV-19/Scotland/QEUH-9DBF44/2020\|EPI_ISL_575338\|2 | ···················· | ···················· | ···················· | ···················· | ···················· | ···················· |
| L | 324 | hCoV-19/Scotland/QEUH-9DA631/2020\|EPI_ISL_575339\|2 | ···················· | ···················· | ···················· | ···················· | ···················· | ···················· |
| L | 325 | hCoV-19/Scotland/QEUH-9DA6B9/2020\|EPI_ISL_575340\|2 | ···················· | ···················· | ···················· | ···················· | ···················· | ···················· |
| L | 326 | hCoV-19/Scotland/QEUH-9DBEA1/2020\|EPI_ISL_575341\|2 | ···················· | ···················· | ···················· | ···················· | ···················· | ···················· |
| L | 327 | hCoV-19/Scotland/QEUH-9DA9B0/2020\|EPI_ISL_575342\|2 | ···················· | ···················· | ···················· | ···················· | ···················· | ···················· |
| L | 328 | hCoV-19/Scotland/QEUH-9DA9ED/2020\|EPI_ISL_575343\|2 | ···················· | ···················· | ···················· | ···················· | ···················· | ···················· |
| L | 329 | hCoV-19/Scotland/QEUH-9DC1C5/2020\|EPI_ISL_575344\|2 | ···················· | ···················· | ···················· | ···················· | ···················· | ···················· |
| L | 330 | hCoV-19/Scotland/QEUH-9DBFBD/2020\|EPI_ISL_575345\|2 | ···················· | ···················· | ···················· | ···················· | ···················· | ···················· |
| L | 331 | hCoV-19/Scotland/QEUH-9DA9CF/2020\|EPI_ISL_575346\|2 | ···················· | ···················· | ···················· | ···················· | ···················· | ···················· |

| | | | | | | | | |
|---|---|---|---|---|---|---|---|---|
| L | 332 | hCoV-19/Scotland/QEUH-9DA5DA/2020\|EPI_ISL_575348\|2 | ···················· | ···················· | ····················· | ···················· | ···················· | ···················· |
| L | 333 | hCoV-19/Scotland/QEUH-9DC09B/2020\|EPI_ISL_575349\|2 | ···················· | ···················· | ····················· | ···················· | ···················· | ···················· |
| L | 334 | hCoV-19/Scotland/QEUH-9DC1B6/2020\|EPI_ISL_575350\|2 | ···················· | ···················· | ····················· | ···················· | ···················· | ···················· |
| L | 335 | hCoV-19/Scotland/QEUH-9DC1A7/2020\|EPI_ISL_575351\|2 | ···················· | ···················· | ····················· | ···················· | ···················· | ···················· |
| L | 336 | hCoV-19/Scotland/QEUH-9DBF71/2020\|EPI_ISL_575352\|2 | ···················· | ···················· | ····················· | ···················· | ···················· | ···················· |
| L | 337 | hCoV-19/Scotland/QEUH-9DC08C/2020\|EPI_ISL_575353\|2 | ···················· | ···················· | ····················· | ···················· | ···················· | ···················· |
| L | 338 | hCoV-19/Scotland/QEUH-9DBFEA/2020\|EPI_ISL_575354\|2 | ···················· | ·A·······A·········· | ····················· | ···················· | ···················· | ···················· |
| L | 339 | hCoV-19/Scotland/QEUH-9DA82C/2020\|EPI_ISL_575355\|2 | ···················· | ···················· | ····················· | ···················· | ···················· | ···················· |
| L | 340 | hCoV-19/Scotland/QEUH-9DA5CB/2020\|EPI_ISL_575356\|2 | ···················· | ···················· | ····················· | ···················· | ···················· | ···················· |
| L | 341 | hCoV-19/Scotland/QEUH-9DAA44/2020\|EPI_ISL_575357\|2 | ···················· | ···················· | ····················· | ···················· | ···················· | ···················· |
| L | 342 | hCoV-19/Scotland/QEUH-9DAA71/2020\|EPI_ISL_575358\|2 | ···················· | ···················· | ····················· | ···················· | ···················· | ···················· |
| L | 343 | hCoV-19/Scotland/QEUH-9DAA17/2020\|EPI_ISL_575359\|2 | ···················· | ···················· | ····················· | ···················· | ···················· | ···················· |
| L | 344 | hCoV-19/Scotland/QEUH-9DBF26/2020\|EPI_ISL_575360\|2 | ···················· | ···················· | ····················· | ···················· | ···················· | ···················· |
| L | 345 | hCoV-19/Scotland/QEUH-9DAA08/2020\|EPI_ISL_575361\|2 | ···················· | ···················· | ····················· | ···················· | ···················· | ···················· |
| L | 346 | hCoV-19/Scotland/QEUH-9DA983/2020\|EPI_ISL_575362\|2 | ···················· | ···················· | ····················· | ···················· | ···················· | ···················· |
| L | 347 | hCoV-19/Scotland/QEUH-9DA9A1/2020\|EPI_ISL_575363\|2 | ···················· | ···················· | ····················· | ···················· | ···················· | ···················· |
| L | 348 | hCoV-19/Scotland/QEUH-9DA992/2020\|EPI_ISL_575364\|2 | ···················· | ···················· | ····················· | ···················· | ···················· | ···················· |
| L | 349 | hCoV-19/Scotland/QEUH-9DC0E6/2020\|EPI_ISL_575365\|2 | ···················· | ···················· | ····················· | ···················· | ···················· | ···················· |
| L | 350 | hCoV-19/Scotland/QEUH-9DC07D/2020\|EPI_ISL_575366\|2 | ···················· | ···················· | ····················· | ···················· | ···················· | ···················· |
| L | 351 | hCoV-19/England/MILK-9E09D5/2020\|EPI_ISL_575367\|20 | ···················· | ···················· | ····················· | ···················· | ···················· | ···················· |
| L | 352 | hCoV-19/Scotland/QEUH-9DBF62/2020\|EPI_ISL_575368\|2 | ···················· | ···················· | ····················· | ···················· | ···················· | ···················· |
| L | 353 | hCoV-19/Scotland/QEUH-9DA9FC/2020\|EPI_ISL_575369\|2 | ···················· | ···················· | ····················· | ···················· | ···················· | ···················· |
| L | 354 | hCoV-19/Scotland/QEUH-9DC1D4/2020\|EPI_ISL_575370\|2 | ···················· | ···················· | ····················· | ···················· | ···················· | ···················· |
| L | 355 | hCoV-19/Scotland/QEUH-9DBF9F/2020\|EPI_ISL_575371\|2 | ···················· | ···················· | ····················· | ···················· | ···················· | ···················· |
| L | 356 | hCoV-19/Scotland/QEUH-9DC0C8/2020\|EPI_ISL_575372\|2 | ···················· | ···················· | ····················· | ···················· | ···················· | ···················· |
| L | 357 | hCoV-19/Scotland/QEUH-9DA877/2020\|EPI_ISL_575373\|2 | ···················· | ···················· | ····················· | ···················· | ···················· | ···················· |
| L | 358 | hCoV-19/Scotland/QEUH-9DAA26/2020\|EPI_ISL_575374\|2 | ···················· | ···················· | ····················· | ···················· | ···················· | ···················· |
| L | 359 | hCoV-19/Scotland/QEUH-9DBF35/2020\|EPI_ISL_575375\|2 | ···················· | ···················· | ····················· | ···················· | ···················· | ···················· |
| L | 360 | hCoV-19/Scotland/QEUH-9DC1E3/2020\|EPI_ISL_575376\|2 | ···················· | ···················· | ····················· | ···················· | ···················· | ···················· |
| L | 361 | hCoV-19/Scotland/QEUH-9DA83B/2020\|EPI_ISL_575377\|2 | ···················· | ···················· | ····················· | ···················· | ···················· | ···················· |
| L | 362 | hCoV-19/Scotland/QEUH-9DC0AA/2020\|EPI_ISL_575378\|2 | ···················· | ···················· | ····················· | ···················· | ···················· | ···················· |
| L | 363 | hCoV-19/England/ALDP-9D8A82/2020\|EPI_ISL_576646\|20 | ···················· | ···················· | ····················· | ···················· | ···················· | ···················· |
| L | 364 | hCoV-19/England/ALDP-9D8A91/2020\|EPI_ISL_576647\|20 | ···················· | ···················· | ····················· | ···················· | ···················· | ···················· |
| L | 365 | hCoV-19/England/ALDP-9D8AFB/2020\|EPI_ISL_576648\|20 | ···················· | ···················· | ····················· | ···················· | ···················· | ···················· |
| L | 366 | hCoV-19/England/ALDP-9D8B16/2020\|EPI_ISL_576649\|20 | ···················· | ···················· | ····················· | ···················· | ···················· | ···················· |
| L | 367 | hCoV-19/England/ALDP-9D8B34/2020\|EPI_ISL_576650\|20 | ···················· | ···················· | ····················· | ···················· | ···················· | ···················· |
| L | 368 | hCoV-19/England/ALDP-9D8B43/2020\|EPI_ISL_576651\|20 | ···················· | ···················· | ····················· | ···················· | ···················· | ···················· |
| L | 369 | hCoV-19/England/ALDP-9D8B61/2020\|EPI_ISL_576652\|20 | ···················· | ·A·················· | ····················· | ···················· | ···················· | ··A················· |
| L | 370 | hCoV-19/England/ALDP-9D8B70/2020\|EPI_ISL_576653\|20 | ···················· | ···················· | ····················· | ···················· | ···················· | ···················· |
| L | 371 | hCoV-19/England/ALDP-9D8BBC/2020\|EPI_ISL_576654\|20 | ···················· | ···················· | ····················· | ···················· | ···················· | ···················· |
| L | 372 | hCoV-19/England/ALDP-9D8C13/2020\|EPI_ISL_576655\|20 | ···················· | ···················· | ····················· | ···················· | ···················· | ···················· |
| L | 373 | hCoV-19/England/ALDP-9D8C22/2020\|EPI_ISL_576656\|20 | ···················· | ···················· | ····················· | ···················· | ···················· | ···················· |
| L | 374 | hCoV-19/England/ALDP-9D8C31/2020\|EPI_ISL_576657\|20 | ···················· | ·A·················· | ····················· | ···················· | ···················· | ···················· |
| L | 375 | hCoV-19/England/ALDP-9D8C40/2020\|EPI_ISL_576658\|20 | ···················· | ···················· | ····················· | ···················· | ···················· | ···················· |
| L | 376 | hCoV-19/England/ALDP-9D8C5F/2020\|EPI_ISL_576659\|20 | ···················· | ···················· | ····················· | ···················· | ···················· | ···················· |
| L | 377 | hCoV-19/England/ALDP-9D8CAA/2020\|EPI_ISL_576660\|20 | ···················· | ···················· | ····················· | ···················· | ···················· | ···················· |
| L | 378 | hCoV-19/England/ALDP-9D8CD7/2020\|EPI_ISL_576662\|20 | ···················· | ···················· | ····················· | ···················· | ···················· | ···················· |
| L | 379 | hCoV-19/England/ALDP-9D8CF5/2020\|EPI_ISL_576663\|20 | ···················· | ···················· | ····················· | ···················· | ···················· | ···················· |
| L | 380 | hCoV-19/England/ALDP-9D8D10/2020\|EPI_ISL_576664\|20 | ···················· | ···················· | ····················· | ···················· | ···················· | ···················· |
| L | 381 | hCoV-19/England/ALDP-9D8D2F/2020\|EPI_ISL_576665\|20 | ···················· | ···················· | ····················· | ···················· | ···················· | ···················· |
| L | 382 | hCoV-19/England/ALDP-9D8D3E/2020\|EPI_ISL_576666\|20 | ···················· | ···················· | ····················· | ···················· | ···················· | ···················· |
| L | 383 | hCoV-19/England/ALDP-9D8D4D/2020\|EPI_ISL_576667\|20 | ···················· | ···················· | ····················· | ···················· | ···················· | ···················· |
| L | 384 | hCoV-19/England/ALDP-9D8D5C/2020\|EPI_ISL_576668\|20 | ···················· | ···················· | ····················· | ···················· | ···················· | ···················· |
| L | 385 | hCoV-19/England/ALDP-9D8D7A/2020\|EPI_ISL_576669\|20 | ···················· | ···················· | ····················· | ···················· | ···················· | ···················· |
| L | 386 | hCoV-19/England/ALDP-9D8D89/2020\|EPI_ISL_576670\|20 | ···················· | ···················· | ····················· | ···················· | ···················· | ···················· |
| L | 387 | hCoV-19/England/ALDP-9D8DB6/2020\|EPI_ISL_576671\|20 | ···················· | ···················· | ····················· | ···················· | ···················· | ···················· |
| L | 388 | hCoV-19/England/ALDP-9D8DF2/2020\|EPI_ISL_576674\|20 | ···················· | ···················· | ····················· | ···················· | ···················· | ···················· |
| L | 389 | hCoV-19/England/ALDP-9D8E0E/2020\|EPI_ISL_576675\|20 | ···················· | ···················· | ····················· | ···················· | ···················· | ···················· |
| L | 390 | hCoV-19/England/ALDP-9D8E3B/2020\|EPI_ISL_576676\|20 | ···················· | ···················· | ····················· | ···················· | ···················· | ···················· |
| L | 391 | hCoV-19/England/ALDP-9D8EB3/2020\|EPI_ISL_576680\|20 | ···················· | ···················· | ····················· | ···················· | ···················· | ···················· |
| L | 392 | hCoV-19/England/ALDP-9D8EC2/2020\|EPI_ISL_576681\|20 | ···················· | ···················· | ····················· | ···················· | ···················· | ···················· |
| L | 393 | hCoV-19/England/ALDP-9D8EE0/2020\|EPI_ISL_576682\|20 | ···················· | ···················· | ····················· | ···················· | ···················· | ···················· |
| L | 394 | hCoV-19/England/ALDP-9D8F1A/2020\|EPI_ISL_576683\|20 | ···················· | ···················· | ····················· | ···················· | ···················· | ···················· |
| L | 395 | hCoV-19/England/ALDP-9D8F29/2020\|EPI_ISL_576684\|20 | ···················· | ···················· | ····················· | ···················· | ···················· | ···················· |
| L | 396 | hCoV-19/England/ALDP-9D8F74/2020\|EPI_ISL_576685\|20 | ···················· | ···················· | ····················· | ···················· | ···················· | ···················· |
| L | 397 | hCoV-19/England/ALDP-9D8F83/2020\|EPI_ISL_576686\|20 | ········T··········· | ···················· | ····················· | ···················· | ···················· | ···················· |
| L | 398 | hCoV-19/England/ALDP-9D8FDE/2020\|EPI_ISL_576687\|20 | ···················· | ···················· | ····················· | ···················· | ···················· | ···················· |
| L | 399 | hCoV-19/England/ALDP-9D8FED/2020\|EPI_ISL_576688\|20 | ···················· | ···················· | ····················· | ···················· | ···················· | ···················· |
| L | 400 | hCoV-19/England/ALDP-9D8FFC/2020\|EPI_ISL_576689\|20 | ···················· | ···················· | ····················· | ···················· | ···················· | ···················· |
| L | 401 | hCoV-19/England/ALDP-9D9007/2020\|EPI_ISL_576690\|20 | ···················· | ···················· | ····················· | ···················· | ···················· | ···················· |
| L | 402 | hCoV-19/England/ALDP-9D9016/2020\|EPI_ISL_576691\|20 | ···················· | ···················· | ····················· | ···················· | ···················· | ···················· |
| L | 403 | hCoV-19/England/ALDP-9D9025/2020\|EPI_ISL_576692\|20 | ···················· | ···················· | ····················· | ···················· | ···················· | ···················· |
| L | 404 | hCoV-19/England/ALDP-9D9034/2020\|EPI_ISL_576693\|20 | ···················· | ···················· | ····················· | ···················· | ···················· | ···················· |
| L | 405 | hCoV-19/England/ALDP-9D9070/2020\|EPI_ISL_576694\|20 | ···················· | ···················· | ····················· | ···················· | ···················· | ···················· |
| L | 406 | hCoV-19/England/ALDP-9D90CB/2020\|EPI_ISL_576696\|20 | ···················· | ···················· | ····················· | ···················· | ···················· | ···················· |
| L | 407 | hCoV-19/England/ALDP-9D90DA/2020\|EPI_ISL_576697\|20 | ···················· | ···················· | ····················· | ···················· | ···················· | ···················· |
| L | 408 | hCoV-19/England/ALDP-9D90E9/2020\|EPI_ISL_576698\|20 | ···················· | ···················· | ····················· | ···················· | ···················· | ···················· |
| L | 409 | hCoV-19/England/ALDP-9D90F8/2020\|EPI_ISL_576699\|20 | ···················· | ···················· | ····················· | ···················· | ···················· | ···················· |
| L | 410 | hCoV-19/Netherlands/UT-EMC-84/2020\|EPI_ISL_577900\| | ···················· | ···················· | ····················· | ···················· | ···················· | ···················· |
| L | 411 | hCoV-19/Netherlands/UT-EMC-85/2020\|EPI_ISL_577901\| | ···················· | ·A·················· | ····················· | ···················· | ···················· | ··A················· |

| | | | | | | | |
|---|---|---|---|---|---|---|---|
| L | 412 hCoV-19/Netherlands/UT-EMC-87/2020\|EPI_ISL_577903\| | ···················· | ···················· | ···················· | ···················· | ···················· | ···················· |
| L | 413 hCoV-19/Netherlands/UT-EMC-88/2020\|EPI_ISL_577904\| | ···················· | ···················· | ···················· | ···················· | ···················· | ···················· |
| L | 414 hCoV-19/Netherlands/UT-EMC-89/2020\|EPI_ISL_577905\| | ···················· | ···················· | ···················· | ···················· | ·····T·············· | ···················· |
| L | 415 hCoV-19/Netherlands/UT-EMC-90/2020\|EPI_ISL_577906\| | ···················· | ···················· | ···················· | ···················· | ···················· | ···················· |
| L | 416 hCoV-19/Netherlands/ZH-EMC-479/2020\|EPI_ISL_577909 | ···················· | ···················· | ···················· | ···················· | ···················· | ···················· |
| L | 417 hCoV-19/Netherlands/ZH-EMC-501/2020\|EPI_ISL_577930 | ···················· | ···················· | ···················· | ···················· | ···················· | ···················· |
| L | 418 hCoV-19/Netherlands/ZH-EMC-547/2020\|EPI_ISL_577975 | ···················· | ···················· | ···················· | ···················· | ···················· | ···················· |
| L | 419 hCoV-19/Netherlands/ZH-EMC-548/2020\|EPI_ISL_577976 | ···················· | ···················· | ···················· | ···················· | ···················· | ···················· |
| L | 420 hCoV-19/Netherlands/ZH-EMC-549/2020\|EPI_ISL_577977 | ···················· | ···················· | ···················· | ···················· | ···················· | ···················· |
| L | 421 hCoV-19/Netherlands/ZH-EMC-550/2020\|EPI_ISL_577978 | ···················· | ···················· | ···················· | ···················· | ···················· | ···················· |
| L | 422 hCoV-19/Netherlands/ZH-EMC-551/2020\|EPI_ISL_577979 | ···················· | ···················· | ···················· | ···················· | ···················· | ···················· |
| L | 423 hCoV-19/Netherlands/ZH-EMC-553/2020\|EPI_ISL_577981 | ···················· | ···················· | ···················· | ···················· | ·····T·············· | ···················· |
| L | 424 hCoV-19/Netherlands/ZH-EMC-554/2020\|EPI_ISL_577982 | ···················· | ···················· | ···················· | ···················· | ·····T·············· | ···················· |
| L | 425 hCoV-19/Netherlands/ZH-EMC-555/2020\|EPI_ISL_577983 | ···················· | ···················· | ···················· | ···················· | ···················· | ···················· |
| L | 426 hCoV-19/Netherlands/ZH-EMC-557/2020\|EPI_ISL_577985 | ···················· | ···················· | ···················· | ···················· | ···················· | ···················· |
| L | 427 hCoV-19/Netherlands/ZH-EMC-561/2020\|EPI_ISL_577989 | ···················· | ···················· | ···················· | ···················· | ···················· | ···················· |
| L | 428 hCoV-19/Netherlands/ZH-EMC-564/2020\|EPI_ISL_577992 | ···················· | ···················· | ···················· | ···················· | ···················· | ···················· |
| L | 429 hCoV-19/Netherlands/ZH-EMC-566/2020\|EPI_ISL_577994 | ···················· | ···················· | ···················· | ···················· | ···················· | ···················· |
| L | 430 hCoV-19/Netherlands/ZH-EMC-568/2020\|EPI_ISL_577996 | ···················· | ···················· | ···················· | ···················· | ···················· | ···················· |
| L | 431 hCoV-19/Netherlands/ZH-EMC-569/2020\|EPI_ISL_577997 | ···················· | ···················· | ···················· | ···················· | ···················· | ···················· |
| L | 432 hCoV-19/Netherlands/ZH-EMC-570/2020\|EPI_ISL_577998 | ···················· | ···················· | ···················· | ···················· | ···················· | ···················· |
| L | 433 hCoV-19/Netherlands/ZH-EMC-571/2020\|EPI_ISL_577999 | ···················· | ···················· | ···················· | ···················· | ···················· | ···················· |
| L | 434 hCoV-19/England/QEUH-9DE9E9/2020\|EPI_ISL_581132\|20 | ···················· | ···················· | ···················· | ···················· | ···················· | ···················· |
| L | 435 hCoV-19/England/QEUH-9DE4AC/2020\|EPI_ISL_581133\|20 | ···················· | ···················· | ···················· | ···················· | ···················· | ···················· |
| L | 436 hCoV-19/Wales/MILK-9E2DD7/2020\|EPI_ISL_581134\|2020 | ···················· | ···················· | ···················· | ···················· | ···················· | ···················· |
| L | 437 hCoV-19/England/MILK-9E2636/2020\|EPI_ISL_581136\|20 | ···················· | ···················· | ···················· | ···················· | ···················· | ···················· |
| L | 438 hCoV-19/England/MILK-9E3073/2020\|EPI_ISL_581137\|20 | ···················· | ···················· | ···················· | ···················· | ···················· | ···················· |
| L | 439 hCoV-19/England/MILK-9E22C9/2020\|EPI_ISL_581138\|20 | ···················· | ···················· | ···················· | ···················· | ···················· | ···················· |
| L | 440 hCoV-19/England/MILK-9E1EF3/2020\|EPI_ISL_581139\|20 | ···················· | ···················· | ···················· | ···················· | ···················· | ···················· |
| L | 441 hCoV-19/England/MILK-9E2988/2020\|EPI_ISL_581140\|20 | ···················· | ···················· | ···················· | ···················· | ···················· | ···················· |
| L | 442 hCoV-19/England/QEUH-9DE5D6/2020\|EPI_ISL_581141\|20 | ···················· | ···················· | ···················· | ···················· | ···················· | ···················· |
| L | 443 hCoV-19/Wales/MILK-9E2A3A/2020\|EPI_ISL_581142\|2020 | ···················· | ···················· | ···················· | ···················· | ···················· | ···················· |
| L | 444 hCoV-19/Wales/MILK-9E2DF5/2020\|EPI_ISL_581143\|2020 | ···················· | ···················· | ···················· | ···················· | ···················· | ···················· |
| L | 445 hCoV-19/England/MILK-9E334D/2020\|EPI_ISL_581144\|20 | ···················· | ···················· | ···················· | ···················· | ···················· | ···················· |
| L | 446 hCoV-19/England/MILK-9E20DE/2020\|EPI_ISL_581145\|20 | ···················· | ···················· | ···················· | ···················· | ···················· | ···················· |
| L | 447 hCoV-19/England/QEUH-9DE5A9/2020\|EPI_ISL_581146\|20 | ···················· | ···················· | ···················· | ···················· | ···················· | ···················· |
| L | 448 hCoV-19/England/MILK-9E235D/2020\|EPI_ISL_581147\|20 | ···················· | ···················· | ···················· | ···················· | ···················· | ···················· |
| L | 449 hCoV-19/England/MILK-9E3055/2020\|EPI_ISL_581148\|20 | ···················· | ···················· | ···················· | ···················· | ···················· | ···················· |
| L | 450 hCoV-19/England/MILK-9E21EA/2020\|EPI_ISL_581151\|20 | ···················· | ·A·················· | ···················· | ···················· | ···················· | ··A················· |
| L | 451 hCoV-19/Wales/MILK-9E1E7B/2020\|EPI_ISL_581152\|2020 | ···················· | ···················· | ···················· | ···················· | ···················· | ···················· |
| L | 452 hCoV-19/Wales/MILK-9E29F1/2020\|EPI_ISL_581153\|2020 | ···················· | ···················· | ···················· | ···················· | ···················· | ···················· |
| L | 453 hCoV-19/England/QEUH-9DEEBD/2020\|EPI_ISL_581154\|20 | ···················· | ···················· | ···················· | ···················· | ···················· | ···················· |
| L | 454 hCoV-19/England/MILK-9E2FB3/2020\|EPI_ISL_581155\|20 | ···················· | ···················· | ···················· | ···················· | ···················· | ···················· |
| L | 455 hCoV-19/England/MILK-9E319E/2020\|EPI_ISL_581156\|20 | ···················· | ···················· | ···················· | ···················· | ···················· | ···················· |
| L | 456 hCoV-19/England/MILK-9E2D40/2020\|EPI_ISL_581157\|20 | ···················· | ···················· | ···················· | ···················· | ···················· | ···················· |
| L | 457 hCoV-19/England/MILK-9E3125/2020\|EPI_ISL_581158\|20 | ···················· | ···················· | ···················· | ···················· | ···················· | ···················· |
| L | 458 hCoV-19/Wales/MILK-9E2144/2020\|EPI_ISL_581159\|2020 | ···················· | ···················· | ···················· | ···················· | ···················· | ···················· |
| L | 459 hCoV-19/England/MILK-9E29A6/2020\|EPI_ISL_581160\|20 | ···················· | ···················· | ···················· | ···················· | ···················· | ···················· |
| L | 460 hCoV-19/Wales/MILK-9E343B/2020\|EPI_ISL_581161\|2020 | ···················· | ···················· | ···················· | ···················· | ···················· | ···················· |
| L | 461 hCoV-19/England/MILK-9E2C34/2020\|EPI_ISL_581162\|20 | ···················· | ··········A········· | ···················· | ···················· | ···················· | ···················· |
| L | 462 hCoV-19/England/MILK-9E2D22/2020\|EPI_ISL_581163\|20 | ···················· | ···················· | ···················· | ···················· | ···················· | ···················· |
| L | 463 hCoV-19/England/MILK-9E2A76/2020\|EPI_ISL_581164\|20 | ···················· | ···················· | ···················· | ···················· | ···················· | ···················· |
| L | 464 hCoV-19/England/MILK-9E33E3/2020\|EPI_ISL_581165\|20 | ···················· | ···················· | ···················· | ···················· | ···················· | ···················· |
| L | 465 hCoV-19/England/QEUH-9DE749/2020\|EPI_ISL_581166\|20 | ···················· | ···················· | ···················· | ···················· | ···················· | ···················· |
| L | 466 hCoV-19/England/MILK-9E2ED4/2020\|EPI_ISL_581167\|20 | ···················· | ···················· | ···················· | ···················· | ···················· | ···················· |
| L | 467 hCoV-19/Wales/MILK-9E3240/2020\|EPI_ISL_581168\|2020 | ···················· | ···················· | ···················· | ···················· | ···················· | ···················· |
| L | 468 hCoV-19/England/MILK-9E1F0F/2020\|EPI_ISL_581169\|20 | ···················· | ···················· | ···················· | ···················· | ···················· | ···················· |
| L | 469 hCoV-19/England/MILK-9E2C16/2020\|EPI_ISL_581170\|20 | ···················· | ···················· | ···················· | ···················· | ···················· | ···················· |
| L | 470 hCoV-19/England/MILK-9E2C07/2020\|EPI_ISL_581171\|20 | ···················· | ···················· | ···················· | ···················· | ···················· | ···················· |
| L | 471 hCoV-19/England/MILK-9E2F59/2020\|EPI_ISL_581172\|20 | ···················· | ·A·················· | ···················· | ···················· | ···················· | ···················· |
| L | 472 hCoV-19/Wales/MILK-9E1E3F/2020\|EPI_ISL_581173\|2020 | ···················· | ···················· | ···················· | ···················· | ···················· | ···················· |
| L | 473 hCoV-19/England/QEUH-9DE600/2020\|EPI_ISL_581174\|20 | ···················· | ···················· | ···················· | ···················· | ···················· | ···················· |
| L | 474 hCoV-19/England/MILK-9E2232/2020\|EPI_ISL_581175\|20 | ···················· | ···················· | ···················· | ···················· | ·····T·············· | ···················· |
| L | 475 hCoV-19/England/MILK-9E2BBF/2020\|EPI_ISL_581199\|20 | ···················· | ···················· | ···················· | ···················· | ···················· | ···················· |
| L | 476 hCoV-19/Canada/MB-NML-1132/2020\|EPI_ISL_582463\|202 | ···················· | ···················· | ···················· | ···················· | ···················· | ···················· |
| L | 477 hCoV-19/Canada/MB-NML-1133/2020\|EPI_ISL_582464\|202 | ···················· | ···················· | ···················· | ···················· | ···················· | ···················· |
| L | 478 hCoV-19/Canada/MB-NML-1134/2020\|EPI_ISL_582465\|202 | ···················· | ···················· | ···················· | ···················· | ···················· | ···················· |
| L | 479 hCoV-19/Canada/MB-NML-1135/2020\|EPI_ISL_582466\|202 | ···················· | ···················· | ···················· | ···················· | ···················· | ···················· |
| L | 480 hCoV-19/Canada/MB-NML-1136/2020\|EPI_ISL_582467\|202 | ···················· | ·A·················· | ···················· | ···················· | ···················· | ··A················· |
| L | 481 hCoV-19/Canada/MB-NML-1138/2020\|EPI_ISL_582468\|202 | ···················· | ···················· | ···················· | ···················· | ···················· | ···················· |
| L | 482 hCoV-19/Canada/MB-NML-1139/2020\|EPI_ISL_582469\|202 | ···················· | ·A·················· | ···················· | ···················· | ···················· | ··A················· |
| L | 483 hCoV-19/Canada/MB-NML-1140/2020\|EPI_ISL_582470\|202 | ···················· | ·A·················· | ···················· | ···················· | ···················· | ··A················· |
| L | 484 hCoV-19/Canada/MB-NML-1141/2020\|EPI_ISL_582471\|202 | ···················· | ·A·················· | ···················· | ···················· | ···················· | ··A················· |
| L | 485 hCoV-19/Canada/MB-NML-1142/2020\|EPI_ISL_582472\|202 | ···················· | ·A·················· | ···················· | ···················· | ···················· | ··A················· |
| L | 486 hCoV-19/Canada/MB-NML-1144/2020\|EPI_ISL_582473\|202 | ···················· | ···················· | ···················· | ···················· | ···················· | ···················· |
| L | 487 hCoV-19/Canada/MB-NML-1146/2020\|EPI_ISL_582474\|202 | ···················· | ···················· | ···················· | ···················· | ···················· | ···················· |
| L | 488 hCoV-19/Canada/MB-NML-1147/2020\|EPI_ISL_582475\|202 | ···················· | ···················· | ···················· | ···················· | ···················· | ···················· |
| L | 489 hCoV-19/Canada/MB-NML-1148/2020\|EPI_ISL_582476\|202 | ···················· | ···················· | ···················· | ···················· | ···················· | ···················· |
| L | 490 hCoV-19/Canada/MB-NML-1150/2020\|EPI_ISL_582477\|202 | ···················· | ···················· | ···················· | ···················· | ···················· | ···················· |
| L | 491 hCoV-19/Canada/MB-NML-1152/2020\|EPI_ISL_582478\|202 | ···················· | ·A·················· | ···················· | ···················· | ···················· | ··A················· |

| | | | | | | | | |
|---|---|---|---|---|---|---|---|---|
| L | 492 | hCoV-19/Canada/MB-NML-1153/2020\|EPI_ISL_582479\|202 | ···················· | ·A·················· | ···················· | ···················· | ···················· | ··A················· |
| L | 493 | hCoV-19/Canada/MB-NML-1154/2020\|EPI_ISL_582480\|202 | ···················· | ·A·················· | ···················· | ···················· | ···················· | ··A················· |
| L | 494 | hCoV-19/Canada/MB-NML-1155/2020\|EPI_ISL_582481\|202 | ···················· | ·A·················· | ···················· | ···················· | ···················· | ··A················· |
| L | 495 | hCoV-19/Canada/MB-NML-1156/2020\|EPI_ISL_582482\|202 | ···················· | ·A·················· | ···················· | ···················· | ···················· | ··A················· |
| L | 496 | hCoV-19/Canada/MB-NML-1157/2020\|EPI_ISL_582483\|202 | ···················· | ·A·················· | ···················· | ···················· | ···················· | ··A················· |
| L | 497 | hCoV-19/Canada/MB-NML-1162/2020\|EPI_ISL_582484\|202 | ···················· | ···················· | ···················· | ···················· | ···················· | ···················· |
| L | 498 | hCoV-19/Canada/MB-NML-1164/2020\|EPI_ISL_582485\|202 | ···················· | ···················· | ···················· | ···················· | ···················· | ···················· |
| L | 499 | hCoV-19/Canada/MB-NML-1165/2020\|EPI_ISL_582486\|202 | ···················· | ···················· | ···················· | ···················· | ···················· | ···················· |
| L | 500 | hCoV-19/Canada/MB-NML-1166/2020\|EPI_ISL_582487\|202 | ···················· | ···················· | ···················· | ···················· | ···················· | ···················· |
| L | 501 | hCoV-19/Canada/MB-NML-1167/2020\|EPI_ISL_582488\|202 | ···················· | ···················· | ···················· | ···················· | ···················· | ···················· |
| L | 502 | hCoV-19/Canada/MB-NML-1168/2020\|EPI_ISL_582489\|202 | ···················· | ·A·················· | ···················· | ···················· | ···················· | ··A················· |
| L | 503 | hCoV-19/Canada/MB-NML-1169/2020\|EPI_ISL_582490\|202 | ···················· | ···················· | ···················· | ···················· | ···················· | ···················· |
| L | 504 | hCoV-19/Canada/MB-NML-1170/2020\|EPI_ISL_582491\|202 | ···················· | ···················· | ···················· | ···················· | ···················· | ···················· |
| L | 505 | hCoV-19/Canada/MB-NML-1171/2020\|EPI_ISL_582492\|202 | ···················· | ·A·················· | ···················· | ···················· | ···················· | ··A················· |
| L | 506 | hCoV-19/Canada/MB-NML-1174/2020\|EPI_ISL_582493\|202 | ···················· | ·A·················· | ···················· | ···················· | ···················· | ···················· |
| L | 507 | hCoV-19/Canada/MB-NML-1176/2020\|EPI_ISL_582494\|202 | ···················· | ·A·················· | ···················· | ···················· | ···················· | ··A················· |
| L | 508 | hCoV-19/Canada/MB-NML-1178/2020\|EPI_ISL_582495\|202 | ···················· | ·A·················· | ···················· | ···················· | ···················· | ··A················· |
| L | 509 | hCoV-19/Canada/MB-NML-1179/2020\|EPI_ISL_582496\|202 | ···················· | ···················· | ···················· | ···················· | ···················· | ···················· |
| L | 510 | hCoV-19/Canada/MB-NML-1180/2020\|EPI_ISL_582497\|202 | ···················· | ···················· | ···················· | ···················· | ···················· | ···················· |
| L | 511 | hCoV-19/Canada/MB-NML-1182/2020\|EPI_ISL_582498\|202 | ···················· | ···················· | ···················· | ···················· | ···················· | ···················· |
| L | 512 | hCoV-19/Canada/MB-NML-1183/2020\|EPI_ISL_582499\|202 | ···················· | ···················· | ···················· | ···················· | ···················· | ···················· |
| L | 513 | hCoV-19/USA/MI-QDX-889/2020\|EPI_ISL_571610\|2020-03 | ···················· | ···················· | ···················· | ···················· | ···················· | ···················· |
| L | 514 | hCoV-19/USA/MI-QDX-890/2020\|EPI_ISL_571611\|2020-03 | ···················· | ···················· | ···················· | ···················· | ···················· | ···················· |
| L | 515 | hCoV-19/USA/MI-QDX-891/2020\|EPI_ISL_571612\|2020-03 | ···················· | ···················· | ···················· | ···················· | ···················· | ···················· |
| L | 516 | hCoV-19/USA/MI-QDX-892/2020\|EPI_ISL_571613\|2020-03 | ···················· | ···················· | ···················· | ···················· | ···················· | ···················· |
| L | 517 | hCoV-19/USA/MI-QDX-893/2020\|EPI_ISL_571614\|2020-03 | ···················· | ···················· | ···················· | ···················· | ···················· | ···················· |
| L | 518 | hCoV-19/USA/MI-QDX-894/2020\|EPI_ISL_571615\|2020-03 | ···················· | ···················· | ···················· | ···················· | ···················· | ···················· |
| L | 519 | hCoV-19/USA/MI-QDX-895/2020\|EPI_ISL_571616\|2020-03 | ···················· | ···················· | ···················· | ···················· | ···················· | ···················· |
| L | 520 | hCoV-19/USA/MI-QDX-896/2020\|EPI_ISL_571617\|2020-03 | ···················· | ···················· | ···················· | ···················· | ···················· | ···················· |
| L | 521 | hCoV-19/USA/MI-QDX-897/2020\|EPI_ISL_571618\|2020-03 | ···················· | ···················· | ···················· | ···················· | ···················· | ···················· |
| L | 522 | hCoV-19/USA/MI-QDX-898/2020\|EPI_ISL_571619\|2020-03 | ···················· | ···················· | ···················· | ···················· | ···················· | ···················· |
| L | 523 | hCoV-19/USA/MI-QDX-899/2020\|EPI_ISL_571620\|2020-03 | ···················· | ···················· | ···················· | ···················· | ···················· | ···················· |
| L | 524 | hCoV-19/USA/MI-QDX-900/2020\|EPI_ISL_571621\|2020-03 | ···················· | ···················· | ···················· | ···················· | ···················· | ···················· |
| L | 525 | hCoV-19/USA/MI-QDX-901/2020\|EPI_ISL_571622\|2020-03 | ···················· | ···················· | ···················· | ···················· | ···················· | ···················· |
| L | 526 | hCoV-19/USA/MI-QDX-902/2020\|EPI_ISL_571623\|2020-03 | ···················· | ···················· | ···················· | ···················· | ···················· | ···················· |
| L | 527 | hCoV-19/USA/MI-QDX-903/2020\|EPI_ISL_571624\|2020-03 | ···················· | ···················· | ···················· | ···················· | ···················· | ···················· |
| L | 528 | hCoV-19/USA/MI-QDX-904/2020\|EPI_ISL_571625\|2020-03 | ···················· | ···················· | ···················· | ···················· | ···················· | ···················· |
| L | 529 | hCoV-19/USA/MI-QDX-907/2020\|EPI_ISL_571626\|2020-03 | ···················· | ···················· | ···················· | ···················· | ···················· | ···················· |
| L | 530 | hCoV-19/USA/MI-QDX-908/2020\|EPI_ISL_571627\|2020-03 | ···················· | ···················· | ···················· | ···················· | ···················· | ···················· |
| L | 531 | hCoV-19/USA/MI-QDX-918/2020\|EPI_ISL_571628\|2020-03 | ···················· | ···················· | ···················· | ···················· | ···················· | ···················· |
| L | 532 | hCoV-19/USA/MI-QDX-919/2020\|EPI_ISL_571629\|2020-03 | ···················· | ···················· | ···················· | ···················· | ···················· | ···················· |
| L | 533 | hCoV-19/USA/MI-QDX-925/2020\|EPI_ISL_571630\|2020-03 | ···················· | ···················· | ···················· | ···················· | ···················· | ···················· |
| L | 534 | hCoV-19/USA/MI-QDX-926/2020\|EPI_ISL_571631\|2020-03 | ···················· | ···················· | ···················· | ···················· | ···················· | ···················· |
| L | 535 | hCoV-19/USA/MI-QDX-927/2020\|EPI_ISL_571632\|2020-03 | ···················· | ···················· | ···················· | ···················· | ···················· | ···················· |
| L | 536 | hCoV-19/USA/MI-QDX-928/2020\|EPI_ISL_571633\|2020-03 | ···················· | ···················· | ···················· | ···················· | ···················· | ···················· |
| L | 537 | hCoV-19/USA/MI-QDX-930/2020\|EPI_ISL_571634\|2020-03 | ···················· | ···················· | ···················· | ···················· | ···················· | ···················· |
| L | 538 | hCoV-19/USA/MI-QDX-931/2020\|EPI_ISL_571635\|2020-03 | ···················· | ···················· | ···················· | ···················· | ···················· | ···················· |
| L | 539 | hCoV-19/USA/MI-QDX-932/2020\|EPI_ISL_571636\|2020-03 | ···················· | ···················· | ···················· | ···················· | ···················· | ···················· |
| L | 540 | hCoV-19/USA/MI-QDX-933/2020\|EPI_ISL_571637\|2020-03 | ···················· | ···················· | ···················· | ···················· | ···················· | ···················· |
| L | 541 | hCoV-19/USA/MI-QDX-934/2020\|EPI_ISL_571638\|2020-03 | ···················· | ···················· | ···················· | ···················· | ···················· | ···················· |
| L | 542 | hCoV-19/USA/MI-QDX-935/2020\|EPI_ISL_571639\|2020-03 | ···················· | ···················· | ···················· | ···················· | ···················· | ···················· |
| L | 543 | hCoV-19/USA/MI-QDX-947/2020\|EPI_ISL_571640\|2020-03 | ···················· | ···················· | ···················· | ···················· | ···················· | ···················· |
| L | 544 | hCoV-19/USA/NJ-QDX-844/2020\|EPI_ISL_571641\|2020-03 | ···················· | ···················· | ···················· | ···················· | ···················· | ···················· |
| L | 545 | hCoV-19/USA/NJ-QDX-849/2020\|EPI_ISL_571642\|2020-03 | ···················· | ···················· | ···················· | ···················· | ···················· | ···················· |
| L | 546 | hCoV-19/USA/NJ-QDX-852/2020\|EPI_ISL_571643\|2020-03 | ···················· | ···················· | ···················· | ···················· | ···················· | ···················· |
| L | 547 | hCoV-19/USA/NJ-QDX-853/2020\|EPI_ISL_571644\|2020-03 | ···················· | ···················· | ···················· | ···················· | ···················· | ···················· |
| L | 548 | hCoV-19/USA/NJ-QDX-866/2020\|EPI_ISL_571645\|2020-03 | ···················· | ···················· | ···················· | ···················· | ···················· | ···················· |
| L | 549 | hCoV-19/USA/NJ-QDX-871/2020\|EPI_ISL_571646\|2020-03 | ···················· | ···················· | ···················· | ···················· | ···················· | ···················· |
| L | 550 | hCoV-19/USA/NY-QDX-833/2020\|EPI_ISL_571647\|2020-03 | ···················· | ···················· | ···················· | ···················· | ···················· | ···················· |
| L | 551 | hCoV-19/USA/NY-QDX-837/2020\|EPI_ISL_571648\|2020-03 | ···················· | ···················· | ···················· | ···················· | ···················· | ···················· |
| L | 552 | hCoV-19/USA/NY-QDX-838/2020\|EPI_ISL_571649\|2020-03 | ···················· | ···················· | ···················· | ···················· | ···················· | ···················· |
| L | 553 | hCoV-19/USA/NY-QDX-839/2020\|EPI_ISL_571650\|2020-03 | ···················· | ···················· | ···················· | ···················· | ···················· | ···················· |
| L | 554 | hCoV-19/USA/NY-QDX-840/2020\|EPI_ISL_571651\|2020-03 | ···················· | ···················· | ···················· | ···················· | ···················· | ···················· |
| L | 555 | hCoV-19/USA/NY-QDX-841/2020\|EPI_ISL_571652\|2020-03 | ···················· | ···················· | ···················· | ···················· | ···················· | ···················· |
| L | 556 | hCoV-19/USA/NY-QDX-842/2020\|EPI_ISL_571653\|2020-03 | ···················· | ···················· | ···················· | ···················· | ···················· | ···················· |
| L | 557 | hCoV-19/USA/NY-QDX-843/2020\|EPI_ISL_571654\|2020-03 | ···················· | ···················· | ···················· | ···················· | ···················· | ···················· |
| L | 558 | hCoV-19/USA/NY-QDX-845/2020\|EPI_ISL_571655\|2020-03 | ···················· | ···················· | ···················· | ···················· | ···················· | ···················· |
| L | 559 | hCoV-19/USA/NY-QDX-846/2020\|EPI_ISL_571656\|2020-03 | ···················· | ···················· | ···················· | ···················· | ···················· | ···················· |
| L | 560 | hCoV-19/USA/NY-QDX-847/2020\|EPI_ISL_571657\|2020-03 | ···················· | ···················· | ···················· | ···················· | ···················· | ···················· |
| L | 561 | hCoV-19/USA/NY-QDX-848/2020\|EPI_ISL_571658\|2020-03 | ···················· | ···················· | ···················· | ···················· | ···················· | ···················· |
| L | 562 | hCoV-19/USA/NY-QDX-850/2020\|EPI_ISL_571659\|2020-03 | ···················· | ···················· | ···················· | ···················· | ···················· | ···················· |
| L | 563 | hCoV-19/USA/NY-QDX-851/2020\|EPI_ISL_571660\|2020-03 | ···················· | ···················· | ···················· | ···················· | ···················· | ···················· |
| L | 564 | hCoV-19/USA/NY-QDX-854/2020\|EPI_ISL_571661\|2020-03 | ···················· | ···················· | ···················· | ···················· | ···················· | ···················· |
| L | 565 | hCoV-19/USA/NY-QDX-855/2020\|EPI_ISL_571662\|2020-03 | ···················· | ···················· | ···················· | ···················· | ···················· | ···················· |
| L | 566 | hCoV-19/USA/NY-QDX-856/2020\|EPI_ISL_571663\|2020-03 | ···················· | ···················· | ···················· | ···················· | ···················· | ···················· |
| L | 567 | hCoV-19/USA/NY-QDX-857/2020\|EPI_ISL_571664\|2020-03 | ···················· | ···················· | ···················· | ···················· | ···················· | ···················· |
| L | 568 | hCoV-19/USA/NY-QDX-858/2020\|EPI_ISL_571665\|2020-03 | ···················· | ···················· | ···················· | ···················· | ···················· | ···················· |
| L | 569 | hCoV-19/USA/NY-QDX-859/2020\|EPI_ISL_571666\|2020-03 | ···················· | ···················· | ···················· | ···················· | ···················· | ···················· |
| L | 570 | hCoV-19/USA/NY-QDX-860/2020\|EPI_ISL_571667\|2020-03 | ···················· | ···················· | ···················· | ···················· | ···················· | ···················· |
| L | 571 | hCoV-19/USA/NY-QDX-861/2020\|EPI_ISL_571668\|2020-03 | ···················· | ···················· | ···················· | ···················· | ···················· | ···················· |

L 572 hCoV-19/USA/NY-QDX-862/2020|EPI_ISL_571669|2020-03 ···················· ···················· ····················· ···················· ···················· ····················
L 573 hCoV-19/USA/NY-QDX-863/2020|EPI_ISL_571670|2020-03 ···················· ···················· ····················· ···················· ···················· ····················
L 574 hCoV-19/USA/NY-QDX-864/2020|EPI_ISL_571671|2020-03 ···················· ···················· ····················· ···················· ···················· ····················
L 575 hCoV-19/USA/NY-QDX-876/2020|EPI_ISL_571680|2020-03 ···················· ···················· ····················· ···················· ···················· ····················
L 576 hCoV-19/USA/NY-QDX-880/2020|EPI_ISL_571681|2020-03 ···················· ···················· ····················· ···················· ···················· ····················
L 577 hCoV-19/USA/NY-QDX-881/2020|EPI_ISL_571682|2020-03 ···················· ···················· ····················· ···················· ···················· ····················
L 578 hCoV-19/USA/NY-QDX-882/2020|EPI_ISL_571683|2020-03 ···················· ···················· ····················· ···················· ···················· ····················
L 579 hCoV-19/USA/NY-QDX-883/2020|EPI_ISL_571684|2020-03 ···················· ···················· ····················· ···················· ···················· ····················
L 580 hCoV-19/USA/OH-QDX-906/2020|EPI_ISL_571685|2020-03 ···················· ···················· ····················· ···················· ···················· ····················
L 581 hCoV-19/USA/OH-QDX-914/2020|EPI_ISL_571686|2020-03 ···················· ···················· ····················· ···················· ···················· ····················
L 582 hCoV-19/USA/TX-QDX-834/2020|EPI_ISL_571687|2020-03 ···················· ···················· ····················· ···················· ···················· ····················
L 583 hCoV-19/USA/TX-QDX-835/2020|EPI_ISL_571688|2020-03 ···················· ···················· ····················· ···················· ···················· ····················
L 584 hCoV-19/USA/TX-QDX-878/2020|EPI_ISL_571689|2020-03 ···················· ···················· ····················· ···················· ···················· ····················
L 585 hCoV-19/USA/WA-QDX-917/2020|EPI_ISL_571691|2020-03 ···················· ···················· ····················· ···················· ···················· ····················
L 586 hCoV-19/USA/AZ-QDX-1054/2020|EPI_ISL_571692|2020-0 ···················· ···················· ····················· ···················· ···················· ····················
L 588 hCoV-19/England/ALDP-9CC4A1/2020|EPI_ISL_572941|20 ···················· ···················· ····················· ···················· ···················· ····················
L 590 hCoV-19/England/NOTT-113889/2020|EPI_ISL_572943|20 ···················· ·A·················· ····················· ···················· ···················· ··A·················
L 591 hCoV-19/England/NOTT-113968/2020|EPI_ISL_572944|20 ···················· ·A·················· ····················· ···················· ···················· ··A·················
L 592 hCoV-19/England/QEUH-9B3A9C/2020|EPI_ISL_572945|20 ···················· ···················· ····················· ···················· ····T··············· ····················
L 593 hCoV-19/England/QEUH-9B3EEB/2020|EPI_ISL_572946|20 ···················· ···················· ····················· ···················· ···················· ····················
L 594 hCoV-19/England/QEUH-9BA2AC/2020|EPI_ISL_572947|20 ···················· ···················· ····················· ···················· ···················· ····················
L 595 hCoV-19/England/SHEF-CE059/2020|EPI_ISL_572948|202 ···················· ·A·················· ····················· ···················· ···················· ··A·················
L 596 hCoV-19/England/SHEF-D23FF/2020|EPI_ISL_572949|202 ···················· ·A·················· ····················· ···················· ···················· ··A·················
L 597 hCoV-19/Wales/MILK-9C6F97/2020|EPI_ISL_572950|2020 ···················· ···················· ····················· ···················· ···················· ····················
L 598 hCoV-19/Wales/PHWC-16B864/2020|EPI_ISL_572951|2020 ···················· ···················· ····················· ···················· ···················· ····················
L 600 hCoV-19/England/QEUH-9AECBC/2020|EPI_ISL_572957|20 ···················· ···················· ····················· ···················· ···················· ····················
L 601 hCoV-19/England/ALDP-9B5D82/2020|EPI_ISL_572958|20 ···················· ···················· ····················· ···················· ···················· ····················
L 602 hCoV-19/England/QEUH-9B3FF7/2020|EPI_ISL_572959|20 ···················· ···················· ····················· ···················· ···················· ····················
L 603 hCoV-19/England/QEUH-9B9F97/2020|EPI_ISL_572960|20 ···················· ···················· ····················· ···················· ···················· ····················
L 604 hCoV-19/England/SHEF-C33A6/2020|EPI_ISL_572961|202 ···················· ···················· ····················· ···················· ···················· ····················
L 605 hCoV-19/England/ALDP-9B5CA3/2020|EPI_ISL_572962|20 ···················· ···················· ····················· ···················· ···················· ····················
L 606 hCoV-19/England/ALDP-9B5CD0/2020|EPI_ISL_572963|20 ···················· ···················· ····················· ···················· ···················· ····················
L 607 hCoV-19/England/ALDP-9B5D0A/2020|EPI_ISL_572964|20 ···················· ···················· ····················· ···················· ···················· ····················
L 608 hCoV-19/England/ALDP-9B5D19/2020|EPI_ISL_572965|20 ···················· ···················· ····················· ···················· ···················· ····················
L 609 hCoV-19/England/ALDP-9B5D46/2020|EPI_ISL_572966|20 ···················· ···················· ····················· ···················· ···················· ····················
L 610 hCoV-19/England/ALDP-9B5DFB/2020|EPI_ISL_572967|20 ···················· ···················· ····················· ···················· ···················· ····················
L 611 hCoV-19/England/ALDP-9B5E07/2020|EPI_ISL_572968|20 ···················· ···················· ····················· ···················· ···················· ····················
L 612 hCoV-19/England/ALDP-9B5E16/2020|EPI_ISL_572969|20 ···················· ···················· ····················· ···················· ···················· ····················
L 613 hCoV-19/England/ALDP-9B5E43/2020|EPI_ISL_572970|20 ···················· ·A·················· ····················· ···················· ···················· ··A·················
L 614 hCoV-19/England/ALDP-9B5EAD/2020|EPI_ISL_572971|20 ···················· ···················· ····················· ···················· ···················· ····················
L 615 hCoV-19/England/ALDP-9B5ECB/2020|EPI_ISL_572972|20 ···················· ···················· ····················· ···················· ···················· ····················
L 616 hCoV-19/England/ALDP-9B5EE9/2020|EPI_ISL_572973|20 ···················· ···················· ····················· ···················· ···················· ····················
L 617 hCoV-19/England/ALDP-9B5EF8/2020|EPI_ISL_572974|20 ···················· ···················· ····················· ···················· ···················· ····················
L 618 hCoV-19/England/ALDP-9B5F04/2020|EPI_ISL_572975|20 ···················· ···················· ····················· ···················· ···················· ····················
L 619 hCoV-19/England/ALDP-9B5F13/2020|EPI_ISL_572976|20 ···················· ···················· ····················· ···················· ···················· ····················
L 620 hCoV-19/England/ALDP-9B5F22/2020|EPI_ISL_572977|20 ···················· ···················· ····················· ···················· ······T············· ····················
L 621 hCoV-19/England/ALDP-9B5F5F/2020|EPI_ISL_572978|20 ···················· ···················· ····················· ···················· ······T············· ····················
L 622 hCoV-19/England/ALDP-9B5F6E/2020|EPI_ISL_572979|20 ···················· ···················· ····················· ···················· ···················· ····················
L 623 hCoV-19/England/ALDP-9B5F9B/2020|EPI_ISL_572980|20 ···················· ·A·················· ····················· ···················· ···················· ····················
L 624 hCoV-19/England/ALDP-9B5FE6/2020|EPI_ISL_572981|20 ···················· ·A·················· ····················· ···················· ···················· ····················
L 625 hCoV-19/England/ALDP-9B601F/2020|EPI_ISL_572982|20 ········T··········· ···················· ····················· ···················· ···················· ····················
L 626 hCoV-19/England/ALDP-9B602E/2020|EPI_ISL_572983|20 ···················· ···················· ····················· ···················· ···················· ····················
L 627 hCoV-19/England/ALDP-9B605B/2020|EPI_ISL_572984|20 ···················· ·A·················· ····················· ···················· ···················· ····················
L 628 hCoV-19/England/ALDP-9B606A/2020|EPI_ISL_572985|20 ···················· ·A·················· ····················· ···················· ···················· ····················
L 629 hCoV-19/England/ALDP-9B6097/2020|EPI_ISL_572986|20 ···················· ···················· ····················· ···················· ···················· ····················
L 630 hCoV-19/England/ALDP-9B60D3/2020|EPI_ISL_572987|20 ···················· ·A·······A·········· ····················· ···················· ···················· ····················
L 631 hCoV-19/England/ALDP-9B6149/2020|EPI_ISL_572988|20 ···················· ···················· ····················· ···················· ···················· ····················
L 632 hCoV-19/England/ALDP-9B6185/2020|EPI_ISL_572989|20 ···················· ···················· ····················· ···················· ···················· ····················
L 633 hCoV-19/England/ALDP-9B61A3/2020|EPI_ISL_572990|20 ···················· ···················· ····················· ···················· ···················· ····················
L 634 hCoV-19/England/ALDP-9B61D0/2020|EPI_ISL_572991|20 ···················· ···················· ····················· ···················· ···················· ····················
L 635 hCoV-19/England/ALDP-9B61EF/2020|EPI_ISL_572992|20 ···················· ···················· ····················· ···················· ···················· ····················
L 636 hCoV-19/England/ALDP-9B6255/2020|EPI_ISL_572993|20 ···················· ···················· ····················· ···················· ···················· ····················
L 637 hCoV-19/England/ALDP-9B6264/2020|EPI_ISL_572994|20 ···················· ···················· ····················· ···················· ···················· ····················
L 638 hCoV-19/England/ALDP-9B6273/2020|EPI_ISL_572995|20 ···················· ···················· ····················· ···················· ···················· ····················
L 639 hCoV-19/England/ALDP-9B62CE/2020|EPI_ISL_572996|20 ···················· ···················· ····················· ···················· ···················· ····················
L 640 hCoV-19/England/ALDP-9B62EC/2020|EPI_ISL_572997|20 ···················· ···················· ····················· ···················· ···················· ····················
L 641 hCoV-19/England/ALDP-9B62FB/2020|EPI_ISL_572998|20 ···················· ···················· ····················· ···················· ···················· ····················
L 642 hCoV-19/England/ALDP-9B6325/2020|EPI_ISL_572999|20 ···················· ···················· ····················· ···················· ···················· ····················
L 643 hCoV-19/England/ALDP-9D7959/2020|EPI_ISL_576601|20 ···················· ···················· ····················· ···················· ···················· ····················
L 644 hCoV-19/England/ALDP-9D7968/2020|EPI_ISL_576602|20 ···················· ···················· ····················· ···················· ····T··············· ····················
L 645 hCoV-19/England/ALDP-9D7977/2020|EPI_ISL_576603|20 ···················· ···················· ····················· ···················· ···················· ····················
L 646 hCoV-19/England/ALDP-9D7986/2020|EPI_ISL_576604|20 ···················· ···················· ····················· ···················· ···················· ····················
L 647 hCoV-19/England/ALDP-9D79C2/2020|EPI_ISL_576605|20 ···················· ·A·················· ····················· ···················· ···················· ····················
L 648 hCoV-19/England/ALDP-9D7A1A/2020|EPI_ISL_576606|20 ···················· ···················· ····················· ···················· ···················· ····················
L 649 hCoV-19/England/ALDP-9D7A29/2020|EPI_ISL_576607|20 ···················· ···················· ····················· ···················· ···················· ····················
L 650 hCoV-19/England/ALDP-9D7A47/2020|EPI_ISL_576608|20 ···················· ·A·················· ····················· ···················· ···················· ··A·················
L 651 hCoV-19/England/ALDP-9D7A56/2020|EPI_ISL_576609|20 ···················· ·A·················· ····················· ···················· ···················· ··A·················
L 652 hCoV-19/England/ALDP-9D7A65/2020|EPI_ISL_576610|20 ···················· ···················· ····················· ···················· ···················· ····················
L 653 hCoV-19/England/ALDP-9D7B08/2020|EPI_ISL_576613|20 ···················· ···················· ····················· ···················· ···················· ····················
L 654 hCoV-19/England/ALDP-9D7B26/2020|EPI_ISL_576614|20 ···················· ···················· ····················· ···················· ···················· ····················

| | | | | | | | |
|---|---|---|---|---|---|---|---|
| L | 655 hCoV-19/England/ALDP-9D7B35/2020\|EPI_ISL_576615\|20 | ···················· | ···················· | ···················· | ···················· | ···················· | ···················· |
| L | 656 hCoV-19/England/ALDP-9D7B62/2020\|EPI_ISL_576616\|20 | ···················· | ···················· | ···················· | ···················· | ···················· | ···················· |
| L | 657 hCoV-19/England/ALDP-9D7B9F/2020\|EPI_ISL_576617\|20 | ···················· | ···················· | ···················· | ···················· | ···················· | ···················· |
| L | 658 hCoV-19/England/ALDP-9D7BBD/2020\|EPI_ISL_576618\|20 | ···················· | ···················· | ···················· | ···················· | ···················· | ···················· |
| L | 659 hCoV-19/England/ALDP-9D7C7E/2020\|EPI_ISL_576619\|20 | ···················· | ·A·················· | ···················· | ···················· | ···················· | ··A················· |
| L | 660 hCoV-19/England/ALDP-9D7CAB/2020\|EPI_ISL_576621\|20 | ···················· | ···················· | ···················· | ···················· | ···················· | ···················· |
| L | 661 hCoV-19/Netherlands/ZH-EMC-480/2020\|EPI_ISL_577910 | ···················· | ···················· | ···················· | ···················· | ···················· | ···················· |
| L | 662 hCoV-19/Netherlands/ZH-EMC-481/2020\|EPI_ISL_577911 | ···················· | ···················· | ···················· | ···················· | ···················· | ···················· |
| L | 663 hCoV-19/Netherlands/ZH-EMC-482/2020\|EPI_ISL_577912 | ···················· | ···················· | ···················· | ···················· | ···················· | ···················· |
| L | 664 hCoV-19/Netherlands/ZH-EMC-483/2020\|EPI_ISL_577913 | ···················· | ···················· | ···················· | ···················· | ···················· | ···················· |
| L | 665 hCoV-19/Netherlands/ZH-EMC-484/2020\|EPI_ISL_577914 | ···················· | ···················· | ···················· | ···················· | ···················· | ···················· |
| L | 666 hCoV-19/Netherlands/ZH-EMC-486/2020\|EPI_ISL_577916 | ···················· | ···················· | ···················· | ···················· | ···················· | ···················· |
| L | 667 hCoV-19/Netherlands/ZH-EMC-488/2020\|EPI_ISL_577918 | ···················· | ···················· | ·········C·········· | ···················· | ···················· | ···················· |
| L | 668 hCoV-19/Netherlands/ZH-EMC-489/2020\|EPI_ISL_577919 | ···················· | ···················· | ···················· | ···················· | ···················· | ···················· |
| L | 669 hCoV-19/Netherlands/ZH-EMC-490/2020\|EPI_ISL_577920 | ···················· | ···················· | ···················· | ···················· | ···················· | ···················· |
| L | 670 hCoV-19/Netherlands/ZH-EMC-492/2020\|EPI_ISL_577921 | ···················· | ···················· | ···················· | ···················· | ···················· | ···················· |
| L | 671 hCoV-19/Netherlands/ZH-EMC-493/2020\|EPI_ISL_577922 | ···················· | ···················· | ···················· | ···················· | ···················· | ···················· |
| L | 672 hCoV-19/Netherlands/ZH-EMC-494/2020\|EPI_ISL_577923 | ···················· | ···················· | ···················· | ···················· | ···················· | ···················· |
| L | 673 hCoV-19/Netherlands/ZH-EMC-495/2020\|EPI_ISL_577924 | ···················· | ···················· | ···················· | ···················· | ···················· | ···················· |
| L | 674 hCoV-19/Netherlands/ZH-EMC-496/2020\|EPI_ISL_577925 | ···················· | ···················· | ···················· | ···················· | ···················· | ···················· |
| L | 675 hCoV-19/Netherlands/ZH-EMC-497/2020\|EPI_ISL_577926 | ···················· | ···················· | ···················· | ···················· | ···················· | ···················· |
| L | 676 hCoV-19/Netherlands/ZH-EMC-498/2020\|EPI_ISL_577927 | ···················· | ···················· | ···················· | ···················· | ···················· | ···················· |
| L | 677 hCoV-19/Netherlands/ZH-EMC-500/2020\|EPI_ISL_577929 | ···················· | ···················· | ···················· | ···················· | ···················· | ···················· |
| L | 678 hCoV-19/Netherlands/ZH-EMC-502/2020\|EPI_ISL_577931 | ···················· | ···················· | ···················· | ···················· | ·····T·············· | ···················· |
| L | 679 hCoV-19/Netherlands/ZH-EMC-504/2020\|EPI_ISL_577933 | ···················· | ···················· | ···················· | ···················· | ···················· | ···················· |
| L | 680 hCoV-19/Netherlands/ZH-EMC-505/2020\|EPI_ISL_577934 | ···················· | ···················· | ···················· | ···················· | ·····T·············· | ···················· |
| L | 681 hCoV-19/Netherlands/ZH-EMC-506/2020\|EPI_ISL_577935 | ···················· | ···················· | ···················· | ···················· | ···················· | ···················· |
| L | 682 hCoV-19/Netherlands/ZH-EMC-508/2020\|EPI_ISL_577937 | ···················· | ···················· | ···················· | ···················· | ···················· | ···················· |
| L | 683 hCoV-19/Netherlands/ZH-EMC-509/2020\|EPI_ISL_577938 | ···················· | ···················· | ···················· | ···················· | ···················· | ···················· |
| L | 684 hCoV-19/Netherlands/ZH-EMC-510/2020\|EPI_ISL_577939 | ···················· | ···················· | ···················· | ···················· | ···················· | ···················· |
| L | 685 hCoV-19/Netherlands/ZH-EMC-511/2020\|EPI_ISL_577940 | ···················· | ···················· | ···················· | ···················· | ···················· | ···················· |
| L | 686 hCoV-19/Netherlands/ZH-EMC-512/2020\|EPI_ISL_577941 | ···················· | ···················· | ···················· | ···················· | ···················· | ···················· |
| L | 687 hCoV-19/Netherlands/ZH-EMC-513/2020\|EPI_ISL_577942 | ···················· | ···················· | ···················· | ···················· | ···················· | ···················· |
| L | 688 hCoV-19/Netherlands/ZH-EMC-514/2020\|EPI_ISL_577943 | ···················· | ···················· | ···················· | ···················· | ···················· | ···················· |
| L | 689 hCoV-19/Netherlands/ZH-EMC-516/2020\|EPI_ISL_577944 | ···················· | ···················· | ···················· | ···················· | ···················· | ···················· |
| L | 690 hCoV-19/Netherlands/ZH-EMC-518/2020\|EPI_ISL_577946 | ···················· | ···················· | ···················· | ···················· | ···················· | ···················· |
| L | 691 hCoV-19/Netherlands/ZH-EMC-519/2020\|EPI_ISL_577947 | ···················· | ···················· | ···················· | ···················· | ···················· | ···················· |
| L | 692 hCoV-19/Netherlands/ZH-EMC-520/2020\|EPI_ISL_577948 | ···················· | ···················· | ···················· | ···················· | ···················· | ···················· |
| L | 693 hCoV-19/Netherlands/ZH-EMC-521/2020\|EPI_ISL_577949 | ···················· | ···················· | ···················· | ···················· | ···················· | ···················· |
| L | 694 hCoV-19/Netherlands/ZH-EMC-522/2020\|EPI_ISL_577950 | ···················· | ···················· | ···················· | ···················· | ···················· | ···················· |
| L | 695 hCoV-19/Netherlands/ZH-EMC-523/2020\|EPI_ISL_577951 | ···················· | ···················· | ···················· | ···················· | ···················· | ···················· |
| L | 696 hCoV-19/Netherlands/ZH-EMC-524/2020\|EPI_ISL_577952 | ···················· | ···················· | ···················· | ···················· | ···················· | ···················· |
| L | 697 hCoV-19/Wales/MILK-9E1224/2020\|EPI_ISL_581000\|2020 | ···················· | ···················· | ···················· | ···················· | ···················· | ···················· |
| L | 698 hCoV-19/England/QEUH-9DC787/2020\|EPI_ISL_581001\|20 | ···················· | ···················· | ···················· | ···················· | ···················· | ···················· |
| L | 699 hCoV-19/England/QEUH-9DC602/2020\|EPI_ISL_581002\|20 | ···················· | ···················· | ···················· | ···················· | ···················· | ···················· |
| L | 700 hCoV-19/England/QEUH-9DAE75/2020\|EPI_ISL_581003\|20 | ···················· | ···················· | ···················· | ···················· | ···················· | ···················· |
| L | 701 hCoV-19/England/MILK-9E13B8/2020\|EPI_ISL_581004\|20 | ···················· | ···················· | ···················· | ···················· | ···················· | ···················· |
| L | 702 hCoV-19/England/QEUH-9DC21D/2020\|EPI_ISL_581005\|20 | ···················· | ···················· | ···················· | ···················· | ···················· | ···················· |
| L | 703 hCoV-19/England/QEUH-9DC58D/2020\|EPI_ISL_581006\|20 | ···················· | ···················· | ···················· | ···················· | ···················· | ···················· |
| L | 704 hCoV-19/England/QEUH-9DC5D8/2020\|EPI_ISL_581007\|20 | ···················· | ···················· | ···················· | ···················· | ···················· | ···················· |
| L | 705 hCoV-19/England/MILK-9E0DD9/2020\|EPI_ISL_581008\|20 | ···················· | ···················· | ···················· | ···················· | ···················· | ···················· |
| L | 706 hCoV-19/England/MILK-9E12AC/2020\|EPI_ISL_581009\|20 | ···················· | ···················· | ···················· | ···················· | ···················· | ···················· |
| L | 707 hCoV-19/England/QEUH-9DCF16/2020\|EPI_ISL_581010\|20 | ···················· | ···················· | ···················· | ···················· | ···················· | ···················· |
| L | 708 hCoV-19/England/MILK-9E31CB/2020\|EPI_ISL_581176\|20 | ···················· | ·A·················· | ···················· | ···················· | ···················· | ···················· |
| L | 709 hCoV-19/England/MILK-9E2EA7/2020\|EPI_ISL_581177\|20 | ···················· | ···················· | ···················· | ···················· | ···················· | ···················· |
| L | 710 hCoV-19/Wales/MILK-9E2A85/2020\|EPI_ISL_581178\|2020 | ···················· | ···················· | ···················· | ···················· | ···················· | ···················· |
| L | 711 hCoV-19/England/MILK-9E2171/2020\|EPI_ISL_581179\|20 | ···················· | ···················· | ···················· | ···················· | ···················· | ···················· |
| L | 712 hCoV-19/England/MILK-9E318F/2020\|EPI_ISL_581180\|20 | ···················· | ···················· | ···················· | ···················· | ···················· | ···················· |
| L | 713 hCoV-19/England/MILK-9E1CF9/2020\|EPI_ISL_581181\|20 | ···················· | ···················· | ···················· | ···················· | ···················· | ···················· |
| L | 714 hCoV-19/England/QEUH-9DE7D0/2020\|EPI_ISL_581182\|20 | ···················· | ···················· | ···················· | ···················· | ···················· | ···················· |
| L | 715 hCoV-19/England/QEUH-9DE9CB/2020\|EPI_ISL_581183\|20 | ···················· | ·A·················· | ···················· | ···················· | ···················· | ···················· |
| L | 716 hCoV-19/England/MILK-9E2E6B/2020\|EPI_ISL_581184\|20 | ···················· | ·A·················· | ···················· | ···················· | ···················· | ··A················· |
| L | 717 hCoV-19/England/MILK-9E24C3/2020\|EPI_ISL_581185\|20 | ···················· | ···················· | ···················· | ···················· | ···················· | ···················· |
| L | 718 hCoV-19/England/MILK-9E30CE/2020\|EPI_ISL_581186\|20 | ···················· | ·A·················· | ···················· | ···················· | ···················· | ··A················· |
| L | 719 hCoV-19/England/MILK-9E229C/2020\|EPI_ISL_581187\|20 | ···················· | ···················· | ···················· | ···················· | ···················· | ···················· |
| L | 720 hCoV-19/Wales/MILK-9E21AE/2020\|EPI_ISL_581188\|2020 | ···················· | ···················· | ···················· | ···················· | ···················· | ···················· |
| L | 721 hCoV-19/England/QEUH-9DE828/2020\|EPI_ISL_581189\|20 | ···················· | ·A·················· | ···················· | ···················· | ···················· | ··A················· |
| L | 722 hCoV-19/England/MILK-9E22BA/2020\|EPI_ISL_581190\|20 | ···················· | ···················· | ···················· | ···················· | ···················· | ···················· |
| L | 723 hCoV-19/England/MILK-9E337A/2020\|EPI_ISL_581192\|20 | ···················· | ···················· | ···················· | ···················· | ···················· | ···················· |
| L | 724 hCoV-19/England/QEUH-9DEEEA/2020\|EPI_ISL_581193\|20 | ···················· | ···················· | ···················· | ···················· | ···················· | ···················· |
| L | 725 hCoV-19/England/QEUH-9DEEAE/2020\|EPI_ISL_581194\|20 | ···················· | ···················· | ···················· | ···················· | ···················· | ···················· |
| L | 726 hCoV-19/England/MILK-9E2F86/2020\|EPI_ISL_581195\|20 | ···················· | ·A·················· | ···················· | ···················· | ···················· | ··A················· |
| L | 727 hCoV-19/England/QEUH-9DEC59/2020\|EPI_ISL_581196\|20 | ···················· | ···················· | ···················· | ···················· | ···················· | ···················· |
| L | 728 hCoV-19/England/MILK-9E2241/2020\|EPI_ISL_581197\|20 | ···················· | ···················· | ···················· | ···················· | ···················· | ···················· |
| L | 729 hCoV-19/Wales/MILK-9E2047/2020\|EPI_ISL_581198\|2020 | ···················· | ···················· | ···················· | ···················· | ···················· | ···················· |
| L | 730 hCoV-19/Canada/MB-NML-656/2020\|EPI_ISL_582300\|2020 | ···················· | ···················· | ···················· | ···················· | ···················· | ···················· |
| L | 731 hCoV-19/Canada/MB-NML-657/2020\|EPI_ISL_582301\|2020 | ···················· | ···················· | ···················· | ···················· | ···················· | ···················· |
| L | 732 hCoV-19/Canada/MB-NML-658/2020\|EPI_ISL_582302\|2020 | ···················· | ···················· | ···················· | ···················· | ···················· | ···················· |
| L | 733 hCoV-19/Canada/MB-NML-766/2020\|EPI_ISL_582303\|2020 | ···················· | ···················· | ···················· | ···················· | ···················· | ···················· |
| L | 734 hCoV-19/Canada/MB-NML-768/2020\|EPI_ISL_582304\|2020 | ···················· | ···················· | ···················· | ···················· | ···················· | ···················· |

| | | | | | | | | |
|---|---|---|---|---|---|---|---|---|
| L | 735 | hCoV-19/Canada/MB-NML-769/2020\|EPI_ISL_582305\|2020 | ···················· | ···················· | ····················· | ···················· | ···················· | ···················· |
| L | 736 | hCoV-19/Canada/MB-NML-779/2020\|EPI_ISL_582306\|2020 | ···················· | ···················· | ····················· | ···················· | ···················· | ···················· |
| L | 737 | hCoV-19/Canada/MB-NML-782/2020\|EPI_ISL_582308\|2020 | ···················· | ···················· | ····················· | ···················· | ···················· | ···················· |
| L | 738 | hCoV-19/Canada/MB-NML-788/2020\|EPI_ISL_582311\|2020 | ···················· | ···················· | ····················· | ···················· | ···················· | ···················· |
| L | 739 | hCoV-19/Canada/MB-NML-789/2020\|EPI_ISL_582312\|2020 | ···················· | ···················· | ····················· | ···················· | ···················· | ···················· |
| L | 740 | hCoV-19/Canada/MB-NML-794/2020\|EPI_ISL_582313\|2020 | ···················· | ···················· | ····················· | ···················· | ···················· | ···················· |
| L | 741 | hCoV-19/Canada/MB-NML-795/2020\|EPI_ISL_582314\|2020 | ···················· | ·····A·············· | ····················· | ···················· | ···················· | ···················· |
| L | 742 | hCoV-19/Canada/MB-NML-796/2020\|EPI_ISL_582315\|2020 | ···················· | ···················· | ····················· | ···················· | ···················· | ···················· |
| L | 743 | hCoV-19/Canada/MB-NML-797/2020\|EPI_ISL_582316\|2020 | ···················· | ···················· | ····················· | ···················· | ···················· | ···················· |
| L | 744 | hCoV-19/Canada/MB-NML-798/2020\|EPI_ISL_582317\|2020 | ···················· | ···················· | ····················· | ···················· | ···················· | ···················· |
| L | 745 | hCoV-19/Canada/MB-NML-808/2020\|EPI_ISL_582318\|2020 | ···················· | ···················· | ····················· | ···················· | ···················· | ···················· |
| L | 746 | hCoV-19/Canada/MB-NML-818/2020\|EPI_ISL_582319\|2020 | ···················· | ···················· | ····················· | ···················· | ···················· | ···················· |
| L | 747 | hCoV-19/Canada/MB-NML-823/2020\|EPI_ISL_582320\|2020 | ···················· | ···················· | ····················· | ···················· | ···················· | ···················· |
| L | 748 | hCoV-19/Canada/MB-NML-824/2020\|EPI_ISL_582321\|2020 | ···················· | ···················· | ····················· | ···················· | ···················· | ···················· |
| L | 749 | hCoV-19/Canada/MB-NML-826/2020\|EPI_ISL_582322\|2020 | ···················· | ···················· | ····················· | ···················· | ···················· | ···················· |
| L | 750 | hCoV-19/Canada/MB-NML-827/2020\|EPI_ISL_582323\|2020 | ···················· | ···················· | ····················· | ···················· | ···················· | ···················· |
| L | 751 | hCoV-19/Canada/MB-NML-828/2020\|EPI_ISL_582324\|2020 | ···················· | ···················· | ····················· | ···················· | ···················· | ···················· |
| L | 752 | hCoV-19/Canada/MB-NML-830/2020\|EPI_ISL_582325\|2020 | ···················· | ···················· | ····················· | ···················· | ···················· | ···················· |
| L | 753 | hCoV-19/Canada/MB-NML-831/2020\|EPI_ISL_582326\|2020 | ···················· | ···················· | ····················· | ···················· | ···················· | ···················· |
| L | 754 | hCoV-19/Canada/MB-NML-832/2020\|EPI_ISL_582327\|2020 | ···················· | ···················· | ····················· | ···················· | ···················· | ···················· |
| L | 755 | hCoV-19/Canada/MB-NML-837/2020\|EPI_ISL_582328\|2020 | ···················· | ···················· | ····················· | ···················· | ···················· | ···················· |
| L | 756 | hCoV-19/Canada/MB-NML-838/2020\|EPI_ISL_582329\|2020 | ···················· | ···················· | ····················· | ···················· | ···················· | ···················· |
| L | 757 | hCoV-19/Canada/MB-NML-839/2020\|EPI_ISL_582330\|2020 | ···················· | ···················· | ····················· | ···················· | ···················· | ···················· |
| L | 758 | hCoV-19/Canada/MB-NML-840/2020\|EPI_ISL_582331\|2020 | ···················· | ···················· | ····················· | ···················· | ···················· | ···················· |
| L | 759 | hCoV-19/Canada/MB-NML-841/2020\|EPI_ISL_582332\|2020 | ···················· | ···················· | ····················· | ···················· | ···················· | ···················· |
| L | 760 | hCoV-19/Canada/MB-NML-843/2020\|EPI_ISL_582333\|2020 | ···················· | ···················· | ····················· | ···················· | ···················· | ···················· |
| L | 761 | hCoV-19/Canada/MB-NML-845/2020\|EPI_ISL_582334\|2020 | ···················· | ···················· | ····················· | ···················· | ···················· | ···················· |
| L | 762 | hCoV-19/Canada/MB-NML-846/2020\|EPI_ISL_582335\|2020 | ···················· | ···················· | ····················· | ···················· | ···················· | ···················· |
| L | 763 | hCoV-19/Canada/MB-NML-847/2020\|EPI_ISL_582336\|2020 | ···················· | ···················· | ····················· | ···················· | ···················· | ···················· |
| L | 764 | hCoV-19/Canada/MB-NML-848/2020\|EPI_ISL_582337\|2020 | ···················· | ···················· | ····················· | ···················· | ···················· | ···················· |
| L | 765 | hCoV-19/Canada/MB-NML-853/2020\|EPI_ISL_582338\|2020 | ···················· | ···················· | ····················· | ···················· | ···················· | ···················· |
| L | 766 | hCoV-19/Canada/MB-NML-854/2020\|EPI_ISL_582339\|2020 | ···················· | ···················· | ····················· | ···················· | ···················· | ···················· |
| L | 767 | hCoV-19/Canada/MB-NML-857/2020\|EPI_ISL_582340\|2020 | ···················· | ···················· | ····················· | ···················· | ···················· | ···················· |
| L | 768 | hCoV-19/Canada/MB-NML-858/2020\|EPI_ISL_582341\|2020 | ···················· | ···················· | ····················· | ···················· | ···················· | ···················· |
| L | 769 | hCoV-19/USA/IL-QDX-831/2020\|EPI_ISL_571600\|2020-03 | ···················· | ···················· | ····················· | ···················· | ···················· | ···················· |
| L | 770 | hCoV-19/USA/IL-QDX-832/2020\|EPI_ISL_571601\|2020-03 | ···················· | ···················· | ····················· | ···················· | ···················· | ···················· |
| L | 771 | hCoV-19/USA/IN-QDX-830/2020\|EPI_ISL_571602\|2020-03 | ···················· | ···················· | ····················· | ···················· | ···················· | ···················· |
| L | 772 | hCoV-19/USA/LA-QDX-836/2020\|EPI_ISL_571603\|2020-03 | ···················· | ···················· | ····················· | ···················· | ···················· | ···················· |
| L | 773 | hCoV-19/USA/LA-QDX-879/2020\|EPI_ISL_571604\|2020-03 | ···················· | ···················· | ····················· | ···················· | ···················· | ···················· |
| L | 774 | hCoV-19/USA/MI-QDX-828/2020\|EPI_ISL_571605\|2020-03 | ···················· | ···················· | ····················· | ···················· | ···················· | ···················· |
| L | 775 | hCoV-19/USA/MI-QDX-885/2020\|EPI_ISL_571606\|2020-03 | ···················· | ···················· | ····················· | ···················· | ···················· | ···················· |
| L | 776 | hCoV-19/USA/MI-QDX-886/2020\|EPI_ISL_571607\|2020-03 | ···················· | ···················· | ····················· | ···················· | ···················· | ···················· |
| L | 777 | hCoV-19/USA/MI-QDX-887/2020\|EPI_ISL_571608\|2020-03 | ···················· | ···················· | ····················· | ···················· | ···················· | ···················· |
| L | 778 | hCoV-19/USA/MI-QDX-888/2020\|EPI_ISL_571609\|2020-03 | ···················· | ···················· | ····················· | ···················· | ···················· | ···················· |
| L | 779 | hCoV-19/USA/NY-QDX-865/2020\|EPI_ISL_571672\|2020-03 | ···················· | ···················· | ····················· | ···················· | ···················· | ···················· |
| L | 780 | hCoV-19/USA/NY-QDX-867/2020\|EPI_ISL_571673\|2020-03 | ···················· | ···················· | ····················· | ···················· | ···················· | ···················· |
| L | 781 | hCoV-19/USA/NY-QDX-868/2020\|EPI_ISL_571674\|2020-03 | ···················· | ···················· | ····················· | ···················· | ···················· | ···················· |
| L | 782 | hCoV-19/USA/NY-QDX-869/2020\|EPI_ISL_571675\|2020-03 | ···················· | ···················· | ····················· | ···················· | ···················· | ···················· |
| L | 783 | hCoV-19/USA/NY-QDX-870/2020\|EPI_ISL_571676\|2020-03 | ···················· | ···················· | ····················· | ···················· | ···················· | ···················· |
| L | 784 | hCoV-19/USA/NY-QDX-872/2020\|EPI_ISL_571677\|2020-03 | ···················· | ···················· | ····················· | ···················· | ···················· | ···················· |
| L | 785 | hCoV-19/USA/NY-QDX-873/2020\|EPI_ISL_571678\|2020-03 | ···················· | ···················· | ····················· | ···················· | ···················· | ···················· |
| L | 786 | hCoV-19/USA/NY-QDX-875/2020\|EPI_ISL_571679\|2020-03 | ···················· | ···················· | ····················· | ···················· | ···················· | ···················· |
| L | 787 | hCoV-19/England/NOTT-1139B3/2020\|EPI_ISL_572938\|20 | ···················· | ·A·················· | ····················· | ···················· | ···················· | ··A················· |
| L | 789 | hCoV-19/USA/UT-UPHL-04307/2020\|EPI_ISL_575137\|2020 | ···················· | ···················· | ····················· | ···················· | ···················· | ···················· |
| L | 790 | hCoV-19/USA/UT-UPHL-04309/2020\|EPI_ISL_575139\|2020 | ···················· | ···················· | ····················· | ···················· | ···················· | ···················· |
| L | 791 | hCoV-19/USA/UT-UPHL-04311/2020\|EPI_ISL_575141\|2020 | ···················· | ···················· | ····················· | ···················· | ···················· | ···················· |
| L | 792 | hCoV-19/USA/UT-UPHL-04312/2020\|EPI_ISL_575142\|2020 | ···················· | ···················· | ····················· | ···················· | ···················· | ···················· |
| L | 793 | hCoV-19/USA/UT-UPHL-04313/2020\|EPI_ISL_575143\|2020 | ···················· | ···················· | ····················· | ···················· | ···················· | ···················· |
| L | 794 | hCoV-19/USA/UT-UPHL-04314/2020\|EPI_ISL_575144\|2020 | ···················· | ···················· | ····················· | ···················· | ···················· | ···················· |
| L | 795 | hCoV-19/USA/UT-UPHL-04318/2020\|EPI_ISL_575147\|2020 | ···················· | ·A·················· | ····················· | ···················· | ···················· | ···················· |
| L | 796 | hCoV-19/USA/UT-UPHL-04319/2020\|EPI_ISL_575148\|2020 | ···················· | ···················· | ····················· | ···················· | ···················· | ···················· |
| L | 797 | hCoV-19/USA/UT-UPHL-04341/2020\|EPI_ISL_575154\|2020 | ···················· | ···················· | ····················· | ···················· | ···················· | ···················· |
| L | 798 | hCoV-19/USA/UT-UPHL-04526/2020\|EPI_ISL_575156\|2020 | ···················· | ···················· | ····················· | ···················· | ···················· | ···················· |
| L | 799 | hCoV-19/USA/UT-UPHL-04528/2020\|EPI_ISL_575157\|2020 | ···················· | ···················· | ····················· | ···················· | ···················· | ···················· |
| L | 800 | hCoV-19/USA/UT-UPHL-04534/2020\|EPI_ISL_575160\|2020 | ···················· | ···················· | ····················· | ···················· | ···················· | ···················· |
| L | 801 | hCoV-19/USA/UT-UPHL-04535/2020\|EPI_ISL_575161\|2020 | ···················· | ···················· | ····················· | ···················· | ···················· | ···················· |
| L | 802 | hCoV-19/USA/UT-UPHL-04536/2020\|EPI_ISL_575162\|2020 | ···················· | ···················· | ····················· | ···················· | ···················· | ···················· |
| L | 803 | hCoV-19/USA/UT-UPHL-04539/2020\|EPI_ISL_575163\|2020 | ···················· | ···················· | ····················· | ···················· | ···················· | ···················· |
| L | 804 | hCoV-19/USA/UT-UPHL-04540/2020\|EPI_ISL_575164\|2020 | ···················· | ···················· | ····················· | ···················· | ···················· | ···················· |
| L | 805 | hCoV-19/USA/UT-UPHL-04541/2020\|EPI_ISL_575165\|2020 | ···················· | ···················· | ····················· | ···················· | ···················· | ···················· |
| L | 806 | hCoV-19/USA/UT-UPHL-04840/2020\|EPI_ISL_575166\|2020 | ···················· | ···················· | ····················· | ···················· | ···················· | ···················· |
| L | 807 | hCoV-19/USA/UT-UPHL-04841/2020\|EPI_ISL_575167\|2020 | ···················· | ···················· | ····················· | ···················· | ···················· | ···················· |
| L | 808 | hCoV-19/USA/UT-UPHL-04849/2020\|EPI_ISL_575173\|2020 | ···················· | ···················· | ····················· | ···················· | ···················· | ···················· |
| L | 809 | hCoV-19/USA/UT-UPHL-04851/2020\|EPI_ISL_575174\|2020 | ···················· | ···················· | ····················· | ···················· | ···················· | ···················· |
| L | 810 | hCoV-19/USA/UT-UPHL-04852/2020\|EPI_ISL_575175\|2020 | ···················· | ···················· | ····················· | ···················· | ···················· | ···················· |
| L | 811 | hCoV-19/USA/UT-UPHL-04856/2020\|EPI_ISL_575177\|2020 | ···················· | ···················· | ····················· | ···················· | ···················· | ···················· |
| L | 812 | hCoV-19/USA/UT-UPHL-04857/2020\|EPI_ISL_575178\|2020 | ···················· | ···················· | ····················· | ···················· | ···················· | ···················· |
| L | 813 | hCoV-19/USA/UT-UPHL-04858/2020\|EPI_ISL_575179\|2020 | ···················· | ···················· | ····················· | ···················· | ···················· | ···················· |
| L | 814 | hCoV-19/USA/UT-UPHL-04859/2020\|EPI_ISL_575180\|2020 | ···················· | ···················· | ····················· | ···················· | ···················· | ···················· |
| L | 815 | hCoV-19/USA/UT-UPHL-04871/2020\|EPI_ISL_575186\|2020 | ···················· | ···················· | ····················· | ···················· | ···················· | ···················· |

| | | | | | | | | |
|---|---|---|---|---|---|---|---|---|
| L | 816 | hCoV-19/USA/UT-UPHL-04887/2020\|EPI_ISL_575188\|2020 | ···················· | ···················· | ···················· | ···················· | ···················· | ···················· |
| L | 817 | hCoV-19/USA/UT-UPHL-04888/2020\|EPI_ISL_575189\|2020 | ···················· | ···················· | ···················· | ···················· | ···················· | ···················· |
| L | 818 | hCoV-19/USA/UT-UPHL-04891/2020\|EPI_ISL_575190\|2020 | ···················· | ···················· | ···················· | ···················· | ···················· | ···················· |
| L | 819 | hCoV-19/USA/UT-UPHL-04895/2020\|EPI_ISL_575191\|2020 | ···················· | ···················· | ···················· | ···················· | ···················· | ···················· |
| L | 820 | hCoV-19/USA/UT-UPHL-04898/2020\|EPI_ISL_575192\|2020 | ···················· | ···················· | ···················· | ···················· | ···················· | ···················· |
| L | 821 | hCoV-19/USA/UT-UPHL-04902/2020\|EPI_ISL_575194\|2020 | ···················· | ···················· | ···················· | ···················· | ···················· | ···················· |
| L | 822 | hCoV-19/USA/UT-UPHL-04904/2020\|EPI_ISL_575196\|2020 | ···················· | ···················· | ···················· | ···················· | ···················· | ···················· |
| L | 823 | hCoV-19/USA/UT-UPHL-04906/2020\|EPI_ISL_575197\|2020 | ···················· | ···················· | ···················· | ···················· | ···················· | ···················· |
| L | 824 | hCoV-19/USA/UT-UPHL-04907/2020\|EPI_ISL_575198\|2020 | ···················· | ···················· | ···················· | ···················· | ···················· | ···················· |
| L | 825 | hCoV-19/USA/UT-UPHL-04909/2020\|EPI_ISL_575199\|2020 | ···················· | ···················· | ···················· | ···················· | ···················· | ···················· |
| L | 826 | hCoV-19/USA/UT-UPHL-04910/2020\|EPI_ISL_575200\|2020 | ···················· | ···················· | ···················· | ···················· | ···················· | ···················· |
| L | 827 | hCoV-19/USA/UT-UPHL-04918/2020\|EPI_ISL_575203\|2020 | ···················· | ···················· | ···················· | ···················· | ···················· | ···················· |
| L | 828 | hCoV-19/USA/UT-UPHL-04919/2020\|EPI_ISL_575204\|2020 | ···················· | ···················· | ···················· | ···················· | ···················· | ···················· |
| L | 829 | hCoV-19/USA/UT-UPHL-04921/2020\|EPI_ISL_575206\|2020 | ···················· | ···················· | ···················· | ···················· | ···················· | ···················· |
| L | 830 | hCoV-19/USA/UT-UPHL-04923/2020\|EPI_ISL_575207\|2020 | ···················· | ···················· | ···················· | ···················· | ···················· | ···················· |
| L | 831 | hCoV-19/USA/UT-UPHL-05518/2020\|EPI_ISL_575210\|2020 | ···················· | ···················· | ···················· | ···················· | ···················· | ···················· |
| L | 832 | hCoV-19/USA/UT-UPHL-05520/2020\|EPI_ISL_575211\|2020 | ···················· | ···················· | ···················· | ···················· | ···················· | ···················· |
| L | 833 | hCoV-19/USA/UT-UPHL-05535/2020\|EPI_ISL_575213\|2020 | ···················· | ···················· | ···················· | ···················· | ···················· | ···················· |
| L | 834 | hCoV-19/USA/UT-UPHL-05540/2020\|EPI_ISL_575214\|2020 | ···················· | ···················· | ···················· | ···················· | ···················· | ···················· |
| L | 835 | hCoV-19/USA/UT-UPHL-05542/2020\|EPI_ISL_575215\|2020 | ···················· | ···················· | ···················· | ···················· | ···················· | ···················· |
| L | 836 | hCoV-19/USA/UT-UPHL-05557/2020\|EPI_ISL_575221\|2020 | ···················· | ···················· | ···················· | ···················· | ···················· | ···················· |
| L | 837 | hCoV-19/USA/UT-UPHL-05559/2020\|EPI_ISL_575222\|2020 | ···················· | ···················· | ···················· | ···················· | ···················· | ···················· |
| L | 838 | hCoV-19/USA/UT-UPHL-05569/2020\|EPI_ISL_575225\|2020 | ···················· | ···················· | ···················· | ···················· | ···················· | ···················· |
| L | 839 | hCoV-19/USA/UT-UPHL-05574/2020\|EPI_ISL_575227\|2020 | ···················· | ···················· | ···················· | ···················· | ···················· | ···················· |
| L | 840 | hCoV-19/USA/UT-UPHL-05575/2020\|EPI_ISL_575228\|2020 | ···················· | ···················· | ···················· | ···················· | ···················· | ···················· |
| L | 841 | hCoV-19/USA/UT-UPHL-05584/2020\|EPI_ISL_575230\|2020 | ···················· | ···················· | ···················· | ···················· | ···················· | ···················· |
| L | 842 | hCoV-19/USA/UT-UPHL-05585/2020\|EPI_ISL_575231\|2020 | ···················· | ···················· | ···················· | ···················· | ···················· | ···················· |
| L | 843 | hCoV-19/USA/UT-UPHL-05586/2020\|EPI_ISL_575232\|2020 | ···················· | ···················· | ···················· | ···················· | ···················· | ···················· |
| L | 844 | hCoV-19/USA/UT-UPHL-05588/2020\|EPI_ISL_575234\|2020 | ···················· | ···················· | ···················· | ···················· | ···················· | ···················· |
| L | 845 | hCoV-19/USA/UT-UPHL-05590/2020\|EPI_ISL_575236\|2020 | ···················· | ···················· | ···················· | ···················· | ···················· | ···················· |
| L | 846 | hCoV-19/USA/UT-UPHL-05591/2020\|EPI_ISL_575237\|2020 | ···················· | ···················· | ···················· | ···················· | ···················· | ···················· |
| L | 847 | hCoV-19/USA/UT-UPHL-2011918/2020\|EPI_ISL_575239\|20 | ···················· | ···················· | ···················· | ···················· | ···················· | ···················· |
| L | 848 | hCoV-19/USA/WA-UW-23510/2020\|EPI_ISL_576468\|2020-0 | ···················· | ·A·················· | ···················· | ···················· | ···················· | ···················· |
| L | 849 | hCoV-19/USA/WA-UW-23511/2020\|EPI_ISL_576469\|2020-0 | ···················· | ···················· | ···················· | ···················· | ···················· | ···················· |
| L | 850 | hCoV-19/USA/WA-UW-23526/2020\|EPI_ISL_576470\|2020-0 | ···················· | ···················· | ···················· | ···················· | ···················· | ···················· |
| L | 851 | hCoV-19/USA/WA-UW-23530/2020\|EPI_ISL_576471\|2020-0 | ···················· | ·A·················· | ···················· | ···················· | ···················· | ···················· |
| L | 852 | hCoV-19/USA/WA-UW-23533/2020\|EPI_ISL_576472\|2020-0 | ···················· | ·A·················· | ···················· | ···················· | ···················· | ···················· |
| L | 853 | hCoV-19/USA/WA-UW-23538/2020\|EPI_ISL_576473\|2020-0 | ···················· | ···················· | ···················· | ···················· | ···················· | ···················· |
| L | 854 | hCoV-19/USA/OR-UW-23549/2020\|EPI_ISL_576474\|2020-0 | ···················· | ···················· | ···················· | ···················· | ···················· | ···················· |
| L | 855 | hCoV-19/USA/WA-UW-23556/2020\|EPI_ISL_576475\|2020-0 | ···················· | ·A·················· | ···················· | ···················· | ···················· | ···················· |
| L | 856 | hCoV-19/USA/WA-UW-23566/2020\|EPI_ISL_576476\|2020-0 | ···················· | ·A·················· | ···················· | ···················· | ···················· | ···················· |
| L | 857 | hCoV-19/USA/WA-UW-23576/2020\|EPI_ISL_576477\|2020-0 | ···················· | ·A·················· | ···················· | ···················· | ···················· | ···················· |
| L | 858 | hCoV-19/USA/WA-UW-23579/2020\|EPI_ISL_576478\|2020-0 | ···················· | ·A·················· | ···················· | ···················· | ···················· | ···················· |
| L | 859 | hCoV-19/USA/WA-UW-23582/2020\|EPI_ISL_576479\|2020-0 | ···················· | ···················· | ···················· | ···················· | ···················· | ···················· |
| L | 860 | hCoV-19/USA/WA-UW-23594/2020\|EPI_ISL_576480\|2020-0 | ···················· | ···················· | ···················· | ···················· | ···················· | ···················· |
| L | 861 | hCoV-19/USA/WA-UW-23596/2020\|EPI_ISL_576481\|2020-0 | ···················· | ·A·················· | ···················· | ···················· | ···················· | ···················· |
| L | 862 | hCoV-19/USA/WA-UW-23597/2020\|EPI_ISL_576482\|2020-0 | ···················· | ···················· | ···················· | ···················· | ···················· | ···················· |
| L | 863 | hCoV-19/USA/WA-UW-23600/2020\|EPI_ISL_576483\|2020-0 | ···················· | ···················· | ···················· | ···················· | ···················· | ···················· |
| L | 864 | hCoV-19/USA/WA-UW-23606/2020\|EPI_ISL_576484\|2020-0 | ···················· | ···················· | ···················· | ···················· | ···················· | ···················· |
| L | 865 | hCoV-19/USA/WA-UW-29527/2020\|EPI_ISL_576487\|2020-0 | ···················· | ·A·················· | ···················· | ···················· | ···················· | ···················· |
| L | 866 | hCoV-19/USA/WA-UW-29528/2020\|EPI_ISL_576488\|2020-0 | ···················· | ·A·················· | ···················· | ···················· | ···················· | ···················· |
| L | 867 | hCoV-19/USA/UN-UW-5231/2020\|EPI_ISL_576489\|2020-04 | ···················· | ···················· | ···················· | ···················· | ···················· | ···················· |
| L | 868 | hCoV-19/USA/UN-UW-5251/2020\|EPI_ISL_576490\|2020-04 | ···················· | ···················· | ···················· | ···················· | ···················· | ···················· |
| L | 869 | hCoV-19/USA/UN-UW-5255/2020\|EPI_ISL_576491\|2020-04 | ···················· | ···················· | ···················· | ···················· | ···················· | ···················· |
| L | 870 | hCoV-19/USA/UN-UW-527/2020\|EPI_ISL_576492\|2020-03- | ···················· | ···················· | ···················· | ···················· | ···················· | ···················· |
| L | 871 | hCoV-19/USA/UN-UW-534/2020\|EPI_ISL_576493\|2020-03- | ···················· | ···················· | ···················· | ···················· | ···················· | ···················· |
| L | 872 | hCoV-19/USA/UN-UW-6996/2020\|EPI_ISL_576494\|2020-03 | ···················· | ···················· | ···················· | ···················· | ···················· | ···················· |
| L | 873 | hCoV-19/USA/UN-UW-6997/2020\|EPI_ISL_576495\|2020-03 | ···················· | ···················· | ···················· | ···················· | ···················· | ···················· |
| L | 874 | hCoV-19/USA/UN-UW-7000/2020\|EPI_ISL_576496\|2020-03 | ···················· | ···················· | ···················· | ···················· | ···················· | ···················· |
| L | 875 | hCoV-19/USA/UN-UW-7002/2020\|EPI_ISL_576497\|2020-03 | ···················· | ···················· | ···················· | ···················· | ···················· | ···················· |
| L | 876 | hCoV-19/USA/UN-UW-7167/2020\|EPI_ISL_576498\|2020-04 | ···················· | ···················· | ···················· | ···················· | ···················· | ···················· |
| L | 877 | hCoV-19/USA/UN-UW-7187/2020\|EPI_ISL_576499\|2020-04 | ···················· | ···················· | ···················· | ···················· | ···················· | ···················· |
| L | 878 | hCoV-19/USA/UN-UW-7204/2020\|EPI_ISL_576501\|2020-04 | ···················· | ···················· | ···················· | ···················· | ···················· | ···················· |
| L | 879 | hCoV-19/USA/UN-UW-7214/2020\|EPI_ISL_576502\|2020-04 | ···················· | ···················· | ···················· | ···················· | ···················· | ···················· |
| L | 880 | hCoV-19/USA/UN-UW-7220/2020\|EPI_ISL_576503\|2020-04 | ···················· | ···················· | ···················· | ···················· | ···················· | ···················· |
| L | 881 | hCoV-19/USA/UN-UW-7227/2020\|EPI_ISL_576504\|2020-04 | ···················· | ···················· | ···················· | ···················· | ···················· | ···················· |
| L | 882 | hCoV-19/USA/UN-UW-7228/2020\|EPI_ISL_576505\|2020-04 | ···················· | ···················· | ···················· | ···················· | ···················· | ···················· |
| L | 883 | hCoV-19/USA/UN-UW-7230/2020\|EPI_ISL_576506\|2020-04 | ···················· | ···················· | ···················· | ···················· | ···················· | ···················· |
| L | 884 | hCoV-19/USA/UN-UW-7231/2020\|EPI_ISL_576507\|2020-04 | ···················· | ···················· | ···················· | ···················· | ···················· | ···················· |
| L | 885 | hCoV-19/USA/UN-UW-7428/2020\|EPI_ISL_576509\|2020-04 | ···················· | ···················· | ···················· | ···················· | ···················· | ···················· |
| L | 886 | hCoV-19/USA/UN-UW-7435/2020\|EPI_ISL_576511\|2020-04 | ···················· | ···················· | ···················· | ···················· | ···················· | ···················· |
| L | 887 | hCoV-19/USA/UN-UW-7455/2020\|EPI_ISL_576512\|2020-04 | ···················· | ···················· | ···················· | ···················· | ···················· | ···················· |
| L | 888 | hCoV-19/USA/UN-UW-7473/2020\|EPI_ISL_576513\|2020-04 | ···················· | ···················· | ···················· | ···················· | ···················· | ···················· |
| L | 889 | hCoV-19/USA/UN-UW-7756/2020\|EPI_ISL_576514\|2020-04 | ···················· | ···················· | ···················· | ···················· | ···················· | ···················· |
| L | 890 | hCoV-19/USA/UN-UW-7770/2020\|EPI_ISL_576515\|2020-04 | ···················· | ···················· | ···················· | ···················· | ···················· | ···················· |
| L | 891 | hCoV-19/USA/UN-UW-7781/2020\|EPI_ISL_576516\|2020-04 | ···················· | ···················· | ···················· | ···················· | ···················· | ···················· |
| L | 892 | hCoV-19/USA/UN-UW-7794/2020\|EPI_ISL_576517\|2020-04 | ···················· | ···················· | ···················· | ···················· | ···················· | ···················· |
| L | 893 | hCoV-19/USA/CA-IGI-0321/2020\|EPI_ISL_576519\|2020-0 | ···················· | ···················· | ···················· | ···················· | ···················· | ···················· |
| L | 894 | hCoV-19/USA/CA-IGI-0324/2020\|EPI_ISL_576520\|2020-0 | ···················· | ···················· | ···················· | ···················· | ···················· | ···················· |
| L | 895 | hCoV-19/USA/CA-IGI-0326/2020\|EPI_ISL_576521\|2020-0 | ···················· | ········A··········· | ···················· | ···················· | ···················· | ···················· |

```
L   896 hCoV-19/USA/CA-IGI-0330/2020|EPI_ISL_576522|2020-0   ····················  ····················  ·····················  ····················  ····················  ····················
L   897 hCoV-19/USA/CA-IGI-0331/2020|EPI_ISL_576523|2020-0   ····················  ····················  ·····················  ····················  ····················  ····················
L   898 hCoV-19/USA/CA-IGI-0333/2020|EPI_ISL_576524|2020-0   ····················  ····················  ·····················  ····················  ····················  ····················
L   899 hCoV-19/USA/CA-IGI-0335/2020|EPI_ISL_576525|2020-0   ····················  ·A··················  ·····················  ····················  ····················  ····················
L   900 hCoV-19/USA/CA-IGI-0336/2020|EPI_ISL_576526|2020-0   ····················  ····················  ·····················  ····················  ····················  ····················
L   901 hCoV-19/USA/CA-IGI-0338/2020|EPI_ISL_576527|2020-0   ····················  ····················  ·····················  ····················  ····················  ····················
L   902 hCoV-19/USA/CA-IGI-0339/2020|EPI_ISL_576528|2020-0   ····················  ····················  ·····················  ····················  ····················  ····················
L   903 hCoV-19/USA/CA-IGI-0340/2020|EPI_ISL_576529|2020-0   ····················  ····················  ·····················  ····················  ····················  ····················
L   904 hCoV-19/USA/CA-IGI-0341/2020|EPI_ISL_576530|2020-0   ····················  ····················  ·····················  ····················  ····················  ····················
L   905 hCoV-19/USA/CA-IGI-0342/2020|EPI_ISL_576531|2020-0   ····················  ····················  ·····················  ····················  ····················  ····················
L   906 hCoV-19/USA/CA-IGI-0344/2020|EPI_ISL_576532|2020-0   ····················  ····················  ·····················  ····················  ····················  ····················
L   907 hCoV-19/USA/CA-IGI-0345/2020|EPI_ISL_576533|2020-0   ····················  ·A··················  ·····················  ····················  ····················  ····················
L   908 hCoV-19/USA/CA-IGI-0346/2020|EPI_ISL_576534|2020-0   ····················  ····················  ·····················  ····················  ····················  ····················
L   909 hCoV-19/USA/CA-IGI-0347/2020|EPI_ISL_576535|2020-0   ····················  ····················  ·····················  ····················  ····················  ····················
L   910 hCoV-19/USA/CA-IGI-0348/2020|EPI_ISL_576536|2020-0   ····················  ····················  ·····················  ····················  ····················  ····················
L   911 hCoV-19/USA/CA-IGI-0351/2020|EPI_ISL_576537|2020-0   ····················  ····················  ·····················  ····················  ····················  ····················
L   912 hCoV-19/USA/CA-IGI-0352/2020|EPI_ISL_576538|2020-0   ····················  ····················  ·····················  ····················  ····················  ····················
L   913 hCoV-19/USA/CA-IGI-0353/2020|EPI_ISL_576539|2020-0   ····················  ·A··················  ·····················  ····················  ····················  ····················
L   914 hCoV-19/USA/CA-IGI-0354/2020|EPI_ISL_576540|2020-0   ····················  ····················  ·····················  ····················  ····················  ····················
L   915 hCoV-19/USA/CA-IGI-0356/2020|EPI_ISL_576541|2020-0   ····················  ····················  ·····················  ····················  ····················  ····················
L   916 hCoV-19/USA/CA-IGI-0359/2020|EPI_ISL_576543|2020-0   ····················  ····················  ·····················  ····················  ····················  ····················
L   917 hCoV-19/USA/CA-IGI-0368/2020|EPI_ISL_576550|2020-0   ····················  ····················  ·····················  ····················  ····················  ····················
L   918 hCoV-19/USA/CA-IGI-0369/2020|EPI_ISL_576551|2020-0   ····················  ····················  ·····················  ····················  ····················  ····················
L   919 hCoV-19/USA/WA-UW-29526/2020|EPI_ISL_576557|2020-0   ····················  ·A··················  ·····················  ····················  ····················  ····················
L   920 hCoV-19/USA/WA-UW-29895/2020|EPI_ISL_576558|2020-1   ····················  ·A··················  ·····················  ····················  ····················  ····················
L   921 hCoV-19/USA/WA-UW-29897/2020|EPI_ISL_576559|2020-1   ····················  ·A··················  ·····················  ····················  ····················  ····················
L   922 hCoV-19/USA/WA-UW-29898/2020|EPI_ISL_576560|2020-1   ····················  ·A··················  ·····················  ····················  ····················  ····················
L   923 hCoV-19/USA/WA-UW-29900/2020|EPI_ISL_576561|2020-1   ····················  ·A··················  ·····················  ····················  ····················  ····················
L   924 hCoV-19/USA/WA-UW-29959/2020|EPI_ISL_576562|2020-1   ····················  ·A··················  ·····················  ····················  ····················  ····················
L   925 hCoV-19/USA/WA-UW-29896/2020|EPI_ISL_576563|2020-1   ····················  ·A··················  ·····················  ····················  ····················  ····················
L   926 hCoV-19/USA/WA-UW-29899/2020|EPI_ISL_576564|2020-1   ····················  ·A··················  ·····················  ····················  ····················  ····················
L   927 hCoV-19/England/ALDP-9D75BF/2020|EPI_ISL_576569|20   ····················  ····················  ·····················  ····················  ····················  ····················
L   928 hCoV-19/England/ALDP-9D75CE/2020|EPI_ISL_576570|20   ····················  ····················  ·····················  ····················  ····················  ····················
L   929 hCoV-19/England/ALDP-9D7607/2020|EPI_ISL_576571|20   ····················  ····················  ·····················  ····················  ····················  ····················
L   930 hCoV-19/England/ALDP-9D7616/2020|EPI_ISL_576572|20   ····················  ····················  ·····················  ····················  ····················  ····················
L   931 hCoV-19/England/ALDP-9D7634/2020|EPI_ISL_576573|20   ····················  ····················  ·····················  ····················  ····················  ····················
L   932 hCoV-19/England/ALDP-9D7643/2020|EPI_ISL_576574|20   ····················  ····················  ·····················  ····················  ····················  ····················
L   933 hCoV-19/England/ALDP-9D7661/2020|EPI_ISL_576575|20   ····················  ····················  ·····················  ····················  ····················  ····················
L   934 hCoV-19/England/ALDP-9D7670/2020|EPI_ISL_576576|20   ····················  ····················  ·····················  ····················  ····················  ····················
L   935 hCoV-19/England/ALDP-9D768F/2020|EPI_ISL_576577|20   ····················  ····················  ·····················  ····················  ····················  ····················
L   936 hCoV-19/England/ALDP-9D76AD/2020|EPI_ISL_576578|20   ····················  ····················  ·····················  ····················  ····················  ····················
L   937 hCoV-19/England/ALDP-9D76BC/2020|EPI_ISL_576579|20   ····················  ····················  ·····················  ····················  ····················  ····················
L   938 hCoV-19/England/ALDP-9D76DA/2020|EPI_ISL_576580|20   ····················  ····················  ·····················  ····················  ····················  ····················
L   939 hCoV-19/England/ALDP-9D76E9/2020|EPI_ISL_576581|20   ····················  ····················  ·····················  ····················  ····················  ····················
L   940 hCoV-19/England/ALDP-9D7713/2020|EPI_ISL_576582|20   ····················  ····················  ·····················  ····················  ····················  ····················
L   941 hCoV-19/England/ALDP-9D7722/2020|EPI_ISL_576583|20   ····················  ····················  ·····················  ····················  ····················  ····················
L   942 hCoV-19/England/ALDP-9D776E/2020|EPI_ISL_576584|20   ····················  ····················  ·····················  ····················  ····················  ····················
L   943 hCoV-19/England/ALDP-9D77C8/2020|EPI_ISL_576585|20   ····················  ····················  ·····················  ····················  ····················  ····················
L   944 hCoV-19/England/ALDP-9D7810/2020|EPI_ISL_576587|20   ····················  ····················  ·····················  ····················  ····················  ····················
L   945 hCoV-19/mink/Netherlands/NB-EMC-44-3/2020|EPI_ISL_   ····················  ····················  ·····················  ····················  ·····T··············  ····················
L   946 hCoV-19/Netherlands/GE-EMC-305/2020|EPI_ISL_577832   ····················  ····················  ·····················  ····················  ····················  ····················
L   947 hCoV-19/Netherlands/LI-EMC-87/2020|EPI_ISL_577833|   ····················  ····················  ·····················  ····················  ····················  ····················
L   948 hCoV-19/Netherlands/LI-EMC-88/2020|EPI_ISL_577834|   ····················  ····················  ·····················  ····················  ····················  ····················
L   949 hCoV-19/Netherlands/LI-EMC-89/2020|EPI_ISL_577835|   ····················  ····················  ·····················  ····················  ····················  ····················
L   950 hCoV-19/Netherlands/NB-EMC-288/2020|EPI_ISL_577837   ····················  ····················  ·····················  ····················  ····················  ····················
L   951 hCoV-19/Netherlands/NB-EMC-289/2020|EPI_ISL_577838   ····················  ····················  ·····················  ····················  ····················  ····················
L   952 hCoV-19/Netherlands/NB-EMC-290/2020|EPI_ISL_577839   ····················  ····················  ·····················  ····················  ····················  ····················
L   953 hCoV-19/Netherlands/NB-EMC-291/2020|EPI_ISL_577840   ····················  ····················  ·····················  ····················  ····················  ····················
L   954 hCoV-19/Netherlands/NB-EMC-292/2020|EPI_ISL_577841   ····················  ····················  ·····················  ····················  ····················  ····················
L   955 hCoV-19/Netherlands/NB-EMC-293/2020|EPI_ISL_577842   ····················  ····················  ·····················  ····················  ····················  ····················
L   956 hCoV-19/Netherlands/NB-EMC-294/2020|EPI_ISL_577843   ····················  ····················  ·····················  ····················  ····················  ····················
L   957 hCoV-19/Netherlands/NB-EMC-295/2020|EPI_ISL_577844   ····················  ·A··················  ·····················  ····················  ····················  ···A················
L   958 hCoV-19/Netherlands/NB-EMC-296/2020|EPI_ISL_577845   ····················  ····················  ·····················  ····················  ·····T··············  ····················
L   959 hCoV-19/Netherlands/NB-EMC-297/2020|EPI_ISL_577846   ····················  ····················  ·····················  ····················  ·····T··············  ····················
L   960 hCoV-19/Netherlands/NH-EMC-51/2020|EPI_ISL_577847|   ····················  ····················  ·····················  ····················  ····················  ····················
L   961 hCoV-19/Netherlands/NH-EMC-52/2020|EPI_ISL_577848|   ····················  ····················  ·····················  ····················  ····················  ····················
L   962 hCoV-19/Netherlands/NH-EMC-54/2020|EPI_ISL_577850|   ····················  ····················  ·····················  ····················  ····················  ····················
L   963 hCoV-19/Netherlands/NH-EMC-55/2020|EPI_ISL_577851|   ····················  ····················  ·····················  ····················  ····················  ····················
L   964 hCoV-19/Netherlands/NH-EMC-56/2020|EPI_ISL_577852|   ····················  ····················  ·····················  ····················  ····················  ····················
L   965 hCoV-19/Netherlands/NH-EMC-57/2020|EPI_ISL_577853|   ····················  ····················  ·····················  ····················  ····················  ····················
L   966 hCoV-19/Netherlands/NH-EMC-58/2020|EPI_ISL_577854|   ····················  ····················  ·····················  ····················  ····················  ····················
L   967 hCoV-19/Netherlands/NH-EMC-59/2020|EPI_ISL_577855|   ····················  ····················  ·····················  ····················  ····················  ····················
L   968 hCoV-19/Netherlands/NH-EMC-60/2020|EPI_ISL_577856|   ····················  ····················  ·····················  ····················  ····················  ····················
L   969 hCoV-19/Netherlands/NH-EMC-61/2020|EPI_ISL_577857|   ····················  ····················  ·····················  ····················  ····················  ····················
L   970 hCoV-19/Netherlands/NH-EMC-62/2020|EPI_ISL_577858|   ····················  ····················  ·····················  ····················  ····················  ····················
L   971 hCoV-19/Netherlands/NH-EMC-63/2020|EPI_ISL_577859|   ····················  ····················  ·····················  ····················  ····················  ····················
L   972 hCoV-19/Netherlands/NH-EMC-65/2020|EPI_ISL_577861|   ····················  ····················  ·····················  ····················  ····················  ····················
L   973 hCoV-19/Netherlands/NH-EMC-67/2020|EPI_ISL_577863|   ····················  ····················  ·····················  ····················  ····················  ····················
L   974 hCoV-19/Netherlands/NH-EMC-68/2020|EPI_ISL_577864|   ····················  ····················  ·····················  ····················  ····················  ····················
L   975 hCoV-19/Netherlands/NH-EMC-69/2020|EPI_ISL_577865|   ····················  ····················  ·····················  ····················  ····················  ····················
```

```
L 976 hCoV-19/Netherlands/NH-EMC-70/2020|EPI_ISL_577866| ···················· ···················· ···················· ···················· ···················· ····················
L 977 hCoV-19/Netherlands/un-EMC-772/2020|EPI_ISL_577868 ···················· ···················· ···················· ···················· ······T············· ····················
L 978 hCoV-19/Netherlands/un-EMC-773/2020|EPI_ISL_577869 ···················· ···················· ···················· ···················· ······T············· ····················
L 979 hCoV-19/Netherlands/un-EMC-774/2020|EPI_ISL_577870 ···················· ···················· ···················· ···················· ······T············· ····················
L 980 hCoV-19/Netherlands/un-EMC-775/2020|EPI_ISL_577871 ···················· ···················· ···················· ···················· ······T············· ····················
L 981 hCoV-19/Netherlands/un-EMC-777/2020|EPI_ISL_577873 ···················· ···················· ···················· ···················· ···················· ····················
L 982 hCoV-19/Netherlands/un-EMC-783/2020|EPI_ISL_577879 ···················· ···················· ···················· ···················· ···················· ····················
L 983 hCoV-19/Netherlands/un-EMC-784/2020|EPI_ISL_577880 ···················· ···················· ···················· ···················· ···················· ····················
L 984 hCoV-19/Netherlands/un-EMC-786/2020|EPI_ISL_577882 ···················· ···················· ···················· ···················· ···················· ····················
L 985 hCoV-19/Netherlands/un-EMC-788/2020|EPI_ISL_577884 ···················· ···················· ···················· ···················· ···················· ····················
L 986 hCoV-19/Netherlands/UT-EMC-70/2020|EPI_ISL_577886| ···················· ···················· ···················· ···················· ···················· ····················
L 987 hCoV-19/Netherlands/UT-EMC-71/2020|EPI_ISL_577887| ···················· ···················· ···················· ···················· ···················· ····················
L 988 hCoV-19/Netherlands/UT-EMC-72/2020|EPI_ISL_577888| ···················· ···················· ···················· ···················· ···················· ····················
L 989 hCoV-19/Netherlands/UT-EMC-73/2020|EPI_ISL_577889| ···················· ···················· ···················· ···················· ···················· ····················
L 990 hCoV-19/Netherlands/UT-EMC-74/2020|EPI_ISL_577890| ···················· ·A·················· ···················· ···················· ···················· ··A·················
L 991 hCoV-19/Netherlands/UT-EMC-75/2020|EPI_ISL_577891| ···················· ·A·················· ···················· ···················· ···················· ··A·················
L 992 hCoV-19/Netherlands/UT-EMC-76/2020|EPI_ISL_577892| ···················· ···················· ···················· ···················· ···················· ····················
L 993 hCoV-19/Netherlands/UT-EMC-77/2020|EPI_ISL_577893| ···················· ·A·················· ···················· ···················· ···················· ··A·················
L 994 hCoV-19/Netherlands/UT-EMC-78/2020|EPI_ISL_577894| ···················· ···················· ···················· ···················· ···················· ····················
L 995 hCoV-19/Netherlands/UT-EMC-79/2020|EPI_ISL_577895| ···················· ···················· ···················· ···················· ···················· ····················
L 996 hCoV-19/Netherlands/UT-EMC-80/2020|EPI_ISL_577896| ···················· ···················· ···················· ···················· ···················· ····················
L 997 hCoV-19/Netherlands/UT-EMC-81/2020|EPI_ISL_577897| ···················· ···················· ···················· ···················· ···················· ····················
L 998 hCoV-19/Netherlands/UT-EMC-82/2020|EPI_ISL_577898| ···················· ···················· ···················· ···················· ···················· ····················
L 999 hCoV-19/Netherlands/UT-EMC-83/2020|EPI_ISL_577899| ···················· ···················· ···················· ···················· ···················· ····················
L 1000 hCoV-19/England/QEUH-9DCDFE/2020|EPI_ISL_581033|20 ···················· ···················· ···················· ···················· ···················· ····················
L 1001 hCoV-19/England/QEUH-9DC7C3/2020|EPI_ISL_581034|20 ···················· ·A·················· ···················· ···················· ···················· ··A·················
L 1002 hCoV-19/England/MILK-9E0D60/2020|EPI_ISL_581035|20 ···················· ···················· ···················· ···················· ······T············· ····················
L 1003 hCoV-19/England/QEUH-9DC73C/2020|EPI_ISL_581036|20 ···················· ···················· ···················· ···················· ···················· ····················
L 1004 hCoV-19/England/QEUH-9DC5F6/2020|EPI_ISL_581037|20 ···················· ···················· ···················· ···················· ···················· ····················
L 1005 hCoV-19/Wales/MILK-9E11FA/2020|EPI_ISL_581038|2020 ···················· ···················· ···················· ···················· ···················· ····················
L 1006 hCoV-19/England/MILK-9E13C7/2020|EPI_ISL_581040|20 ···················· ···················· ···················· ···················· ···················· ····················
L 1007 hCoV-19/England/MILK-9E1312/2020|EPI_ISL_581041|20 ···················· ···················· ···················· ···················· ···················· ····················
L 1008 hCoV-19/England/QEUH-9DCBA9/2020|EPI_ISL_581042|20 ···················· ···················· ···················· ···················· ···················· ····················
L 1009 hCoV-19/England/QEUH-9DC392/2020|EPI_ISL_581043|20 ···················· ·A·················· ···················· ···················· ···················· ··A·················
L 1010 hCoV-19/England/QEUH-9DCFE9/2020|EPI_ISL_581048|20 ···················· ···················· ···················· ···················· ···················· ····················
L 1011 hCoV-19/England/QEUH-9DD2FE/2020|EPI_ISL_581049|20 ···················· ···················· ···················· ···················· ···················· ····················
L 1012 hCoV-19/England/MILK-9E0F00/2020|EPI_ISL_581099|20 ···················· ···················· ···················· ···················· ···················· ····················
L 1013 hCoV-19/Canada/MB-NML-990/2020|EPI_ISL_582364|2020 ···················· ···················· ···················· ···················· ···················· ····················
L 1014 hCoV-19/Canada/MB-NML-991/2020|EPI_ISL_582365|2020 ···················· ···················· ···················· ···················· ···················· ····················
L 1015 hCoV-19/Canada/MB-NML-992/2020|EPI_ISL_582366|2020 ···················· ·A·················· ···················· ···················· ···················· ··A·················
L 1016 hCoV-19/Canada/MB-NML-997/2020|EPI_ISL_582367|2020 ···················· ···················· ···················· ···················· ···················· ····················
L 1017 hCoV-19/Canada/MB-NML-999/2020|EPI_ISL_582368|2020 ···················· ···················· ···················· ···················· ···················· ····················
L 1018 hCoV-19/Canada/MB-NML-1001/2020|EPI_ISL_582369|202 ···················· ···················· ···················· ···················· ···················· ····················
L 1019 hCoV-19/Canada/MB-NML-1002/2020|EPI_ISL_582370|202 ···················· ···················· ···················· ···················· ···················· ····················
L 1020 hCoV-19/Canada/MB-NML-1003/2020|EPI_ISL_582371|202 ···················· ·A·················· ···················· ···················· ···················· ··A·················
L 1021 hCoV-19/Canada/MB-NML-1005/2020|EPI_ISL_582372|202 ···················· ·A·················· ···················· ···················· ···················· ··A·················
L 1022 hCoV-19/Canada/MB-NML-1006/2020|EPI_ISL_582373|202 ···················· ···················· ···················· ···················· ···················· ····················
L 1023 hCoV-19/Canada/MB-NML-1007/2020|EPI_ISL_582374|202 ···················· ···················· ···················· ···················· ···················· ····················
L 1024 hCoV-19/Canada/MB-NML-1012/2020|EPI_ISL_582379|202 ···················· ···················· ···················· ···················· ···················· ····················
L 1025 hCoV-19/USA/CA-QDX-807/2020|EPI_ISL_571500|2020-03 ···················· ···················· ···················· ···················· ···················· ····················
L 1026 hCoV-19/USA/CA-QDX-808/2020|EPI_ISL_571501|2020-03 ···················· ···················· ···················· ···················· ···················· ····················
L 1027 hCoV-19/USA/CA-QDX-809/2020|EPI_ISL_571502|2020-03 ···················· ···················· ···················· ···················· ···················· ····················
L 1028 hCoV-19/USA/CA-QDX-810/2020|EPI_ISL_571503|2020-03 ···················· ···················· ···················· ···················· ···················· ····················
L 1029 hCoV-19/USA/CA-QDX-811/2020|EPI_ISL_571504|2020-03 ···················· ···················· ···················· ···················· ···················· ····················
L 1030 hCoV-19/USA/CA-QDX-814/2020|EPI_ISL_571505|2020-03 ···················· ···················· ···················· ···················· ···················· ····················
L 1031 hCoV-19/USA/CA-QDX-815/2020|EPI_ISL_571506|2020-03 ···················· ···················· ···················· ···················· ···················· ····················
L 1032 hCoV-19/USA/CA-QDX-816/2020|EPI_ISL_571507|2020-03 ···················· ···················· ···················· ···················· ···················· ····················
L 1033 hCoV-19/USA/CA-QDX-817/2020|EPI_ISL_571508|2020-03 ···················· ···················· ···················· ···················· ···················· ····················
L 1034 hCoV-19/USA/CT-QDX-760/2020|EPI_ISL_571509|2020-03 ···················· ···················· ···················· ···················· ···················· ····················
L 1035 hCoV-19/USA/CT-QDX-761/2020|EPI_ISL_571510|2020-03 ···················· ···················A ···················· ···················· ···················· ···················· 1
L 1036 hCoV-19/USA/CT-QDX-762/2020|EPI_ISL_571511|2020-03 ···················· ···················· ···················· ···················· ···················· ····················
L 1037 hCoV-19/USA/CT-QDX-763/2020|EPI_ISL_571512|2020-03 ···················· ···················· ···················· ···················· ···················· ····················
L 1038 hCoV-19/USA/CT-QDX-765/2020|EPI_ISL_571514|2020-03 ···················· ···················· ···················· ···················· ···················· ····················
L 1039 hCoV-19/USA/CT-QDX-766/2020|EPI_ISL_571515|2020-03 ···················· ···················· ···················· ···················· ···················· ····················
L 1040 hCoV-19/USA/CT-QDX-767/2020|EPI_ISL_571516|2020-03 ···················· ···················· ···················· ···················· ···················· ····················
L 1041 hCoV-19/USA/CT-QDX-768/2020|EPI_ISL_571517|2020-03 ···················· ···················· ···················· ···················· ···················· ····················
L 1042 hCoV-19/USA/CT-QDX-769/2020|EPI_ISL_571518|2020-03 ···················· ···················A ···················· ···················· ···················· ···················· 1
L 1043 hCoV-19/USA/FL-QDX-752/2020|EPI_ISL_571519|2020-03 ···················· ···················· ···················· ···················· ···················· ····················
L 1044 hCoV-19/USA/FL-QDX-785/2020|EPI_ISL_571520|2020-03 ···················· ···················· ···················· ···················· ···················· ····················
L 1045 hCoV-19/USA/FL-QDX-786/2020|EPI_ISL_571521|2020-03 ···················· ···················· ···················· ···················· ···················· ····················
L 1046 hCoV-19/USA/FL-QDX-787/2020|EPI_ISL_571522|2020-03 ···················· ···················· ···················· ···················· ···················· ····················
L 1047 hCoV-19/USA/FL-QDX-788/2020|EPI_ISL_571523|2020-03 ···················· ···················· ···················· ···················· ···················· ····················
L 1048 hCoV-19/USA/FL-QDX-790/2020|EPI_ISL_571524|2020-03 ···················· ···················· ···················· ···················· ···················· ····················
L 1049 hCoV-19/USA/FL-QDX-791/2020|EPI_ISL_571525|2020-03 ···················· ···················· ···················· ···················· ···················· ····················
L 1050 hCoV-19/USA/FL-QDX-792/2020|EPI_ISL_571526|2020-03 ···················· ···················· ···················· ···················· ···················· ····················
L 1051 hCoV-19/USA/HI-QDX-758/2020|EPI_ISL_571527|2020-03 ···················· ···················· ···················· ···················· ···················· ····················
L 1052 hCoV-19/USA/HI-QDX-759/2020|EPI_ISL_571528|2020-03 ···················· ···················· ···················· ···················· ···················· ····················
L 1053 hCoV-19/USA/LA-QDX-740/2020|EPI_ISL_571529|2020-03 ···················· ···················· ···················· ···················· ···················· ····················
L 1054 hCoV-19/USA/LA-QDX-741/2020|EPI_ISL_571530|2020-03 ···················· ···················· ···················· ···················· ···················· ····················
L 1055 hCoV-19/USA/LA-QDX-742/2020|EPI_ISL_571531|2020-03 ···················· ···················· ···················· ···················· ···················· ····················
```

| | | | | | | | | |
|---|---|---|---|---|---|---|---|---|
| L | 1056 hCoV-19/USA/MI-QDX-751/2020\|EPI_ISL_571532\|2020-03 | ···················· | ···················· | ···················· | ···················· | ···················· | ···················· | |
| L | 1057 hCoV-19/USA/MI-QDX-754/2020\|EPI_ISL_571533\|2020-03 | ···················· | ···················· | ···················· | ···················· | ···················· | ···················· | |
| L | 1058 hCoV-19/USA/MI-QDX-755/2020\|EPI_ISL_571534\|2020-03 | ···················· | ···················· | ···················· | ···················· | ···················· | ···················· | |
| L | 1059 hCoV-19/USA/MI-QDX-812/2020\|EPI_ISL_571535\|2020-03 | ···················· | ···················· | ···················· | ···················· | ···················· | ···················· | |
| L | 1060 hCoV-19/USA/MI-QDX-819/2020\|EPI_ISL_571536\|2020-03 | ···················· | ···················· | ···················· | ···················· | ···················· | ···················· | |
| L | 1061 hCoV-19/USA/MI-QDX-820/2020\|EPI_ISL_571537\|2020-03 | ···················· | ···················· | ···················· | ···················· | ···················· | ···················· | |
| L | 1062 hCoV-19/USA/MI-QDX-821/2020\|EPI_ISL_571538\|2020-03 | ···················· | ···················· | ···················· | ···················· | ···················· | ···················· | |
| L | 1063 hCoV-19/USA/MI-QDX-822/2020\|EPI_ISL_571539\|2020-03 | ···················· | ···················· | ···················· | ···················· | ···················· | ···················· | |
| L | 1064 hCoV-19/USA/MI-QDX-823/2020\|EPI_ISL_571540\|2020-03 | ···················· | ···················· | ···················· | ···················· | ···················· | ···················· | |
| L | 1065 hCoV-19/USA/MI-QDX-824/2020\|EPI_ISL_571541\|2020-03 | ···················· | ···················· | ···················· | ···················· | ···················· | ···················· | |
| L | 1066 hCoV-19/USA/MI-QDX-826/2020\|EPI_ISL_571543\|2020-03 | ···················· | ···················· | ···················· | ···················· | ···················· | ···················· | |
| L | 1067 hCoV-19/USA/MI-QDX-827/2020\|EPI_ISL_571544\|2020-03 | ···················· | ···················· | ···················· | ···················· | ···················· | ···················· | |
| L | 1068 hCoV-19/USA/MN-QDX-743/2020\|EPI_ISL_571545\|2020-03 | ···················· | ···················· | ···················· | ···················· | ···················· | ···················· | |
| L | 1069 hCoV-19/USA/MN-QDX-744/2020\|EPI_ISL_571546\|2020-03 | ···················· | ···················· | ···················· | ···················· | ···················· | ···················· | |
| L | 1070 hCoV-19/USA/MN-QDX-753/2020\|EPI_ISL_571547\|2020-03 | ···················· | ···················· | ···················· | ···················· | ···················· | ···················· | |
| L | 1071 hCoV-19/USA/MN-QDX-756/2020\|EPI_ISL_571548\|2020-03 | ···················· | ···················· | ···················· | ···················· | ···················· | ···················· | |
| L | 1072 hCoV-19/USA/NV-QDX-757/2020\|EPI_ISL_571549\|2020-03 | ···················· | ···················· | ···················· | ···················· | ···················· | ···················· | |
| L | 1073 hCoV-19/USA/NY-QDX-798/2020\|EPI_ISL_571550\|2020-03 | ···················· | ···················· | ···················· | ···················· | ···················· | ···················· | |
| L | 1074 hCoV-19/USA/OK-QDX-749/2020\|EPI_ISL_571551\|2020-03 | ···················· | ···················· | ···················· | ···················· | ···················· | ···················· | |
| L | 1075 hCoV-19/USA/OK-QDX-750/2020\|EPI_ISL_571552\|2020-03 | ···················· | ···················· | ···················· | ···················· | ···················· | ···················· | |
| L | 1076 hCoV-19/USA/OK-QDX-770/2020\|EPI_ISL_571553\|2020-03 | ···················· | ···················· | ···················· | ···················· | ···················· | ···················· | |
| L | 1077 hCoV-19/USA/OK-QDX-771/2020\|EPI_ISL_571554\|2020-03 | ···················· | ···················· | ···················· | ···················· | ···················· | ···················· | |
| L | 1078 hCoV-19/USA/OK-QDX-772/2020\|EPI_ISL_571555\|2020-03 | ···················· | ···················· | ···················· | ···················· | ···················· | ···················· | |
| L | 1079 hCoV-19/USA/OK-QDX-773/2020\|EPI_ISL_571556\|2020-03 | ···················· | ···················· | ···················· | ···················· | ···················· | ···················· | |
| L | 1080 hCoV-19/USA/TX-QDX-745/2020\|EPI_ISL_571557\|2020-03 | ···················· | ···················· | ···················· | ···················· | ···················· | ···················· | |
| L | 1081 hCoV-19/USA/TX-QDX-746/2020\|EPI_ISL_571558\|2020-03 | ···················· | ···················· | ···················· | ···················· | ···················· | ···················· | |
| L | 1082 hCoV-19/USA/TX-QDX-747/2020\|EPI_ISL_571559\|2020-03 | ···················· | ···················· | ···················· | ···················· | ···················· | ···················· | |
| L | 1083 hCoV-19/USA/TX-QDX-748/2020\|EPI_ISL_571560\|2020-03 | ···················· | ···················· | ···················· | ···················· | ···················· | ···················· | |
| L | 1084 hCoV-19/USA/WA-QDX-813/2020\|EPI_ISL_571561\|2020-03 | ···················· | ···················· | ···················· | ···················· | ···················· | ···················· | |
| L | 1085 hCoV-19/USA/WA-QDX-818/2020\|EPI_ISL_571562\|2020-03 | ···················· | ···················· | ···················· | ···················· | ···················· | ···················· | |
| L | 1086 hCoV-19/USA/WI-QDX-777/2020\|EPI_ISL_571563\|2020-03 | ···················· | ···················· | ···················· | ···················· | ···················· | ···················· | |
| L | 1087 hCoV-19/USA/WI-QDX-778/2020\|EPI_ISL_571564\|2020-03 | ···················· | ···················· | ···················· | ···················· | ···················· | ···················· | |
| L | 1088 hCoV-19/USA/WI-QDX-779/2020\|EPI_ISL_571565\|2020-03 | ···················· | ···················· | ···················· | ···················· | ···················· | ···················· | |
| L | 1089 hCoV-19/USA/WI-QDX-780/2020\|EPI_ISL_571566\|2020-03 | ···················· | ···················· | ···················· | ···················· | ···················· | ···················· | |
| L | 1090 hCoV-19/USA/WI-QDX-782/2020\|EPI_ISL_571567\|2020-03 | ···················· | ···················· | ···················· | ···················· | ···················· | ···················· | |
| L | 1091 hCoV-19/USA/WI-QDX-783/2020\|EPI_ISL_571568\|2020-03 | ···················· | ···················· | ···················· | ···················· | ···················· | ···················· | |
| L | 1092 hCoV-19/USA/WI-QDX-784/2020\|EPI_ISL_571569\|2020-03 | ···················· | ···················· | ···················· | ···················· | ···················· | ···················· | |
| L | 1093 hCoV-19/USA/WI-QDX-789/2020\|EPI_ISL_571570\|2020-03 | ···················· | ···················· | ···················· | ···················· | ···················· | ···················· | |
| L | 1094 hCoV-19/USA/WI-QDX-793/2020\|EPI_ISL_571571\|2020-03 | ···················· | ···················· | ···················· | ···················· | ···················· | ···················· | |
| L | 1095 hCoV-19/USA/AL-QDX-949/2020\|EPI_ISL_571572\|2020-03 | ···················· | ···················· | ···················· | ···················· | ···················· | ···················· | |
| L | 1096 hCoV-19/USA/CA-QDX-1027/2020\|EPI_ISL_571694\|2020-0 | ···················· | ···················· | ···················· | ···················· | ···················· | ···················· | |
| L | 1097 hCoV-19/USA/CA-QDX-1028/2020\|EPI_ISL_571695\|2020-0 | ···················· | ···················· | ···················· | ···················· | ···················· | ···················· | |
| L | 1098 hCoV-19/USA/CA-QDX-1029/2020\|EPI_ISL_571696\|2020-0 | ···················· | ···················· | ···················· | ···················· | ···················· | ···················· | |
| L | 1099 hCoV-19/USA/CA-QDX-1032/2020\|EPI_ISL_571697\|2020-0 | ···················· | ···················· | ···················· | ···················· | ···················· | ···················· | |
| L | 1100 hCoV-19/USA/CA-QDX-1033/2020\|EPI_ISL_571698\|2020-0 | ···················· | ···················· | ···················· | ···················· | ···················· | ···················· | |
| L | 1101 hCoV-19/USA/CA-QDX-1034/2020\|EPI_ISL_571699\|2020-0 | ···················· | ···················· | ···················· | ···················· | ···················· | ···················· | |
| L | 1102 hCoV-19/Scotland/QEUH-9BA42E/2020\|EPI_ISL_572846\|2 | ···················· | ···················· | ···················· | ···················· | ···················· | ···················· | |
| L | 1104 hCoV-19/England/SHEF-D3121/2020\|EPI_ISL_572848\|202 | ···················· | ···················· | ···················· | ···················· | ···················· | ···················· | |
| L | 1105 hCoV-19/England/ALDP-9B5EDA/2020\|EPI_ISL_572849\|20 | ···················· | ···················· | ···················· | ···················· | ···················· | ···················· | |
| L | 1106 hCoV-19/England/ALDP-9BCEA6/2020\|EPI_ISL_572850\|20 | ···················· | ···················· | ···················· | ···················· | ···················· | ···················· | |
| L | 1107 hCoV-19/England/CAMB-1B6044/2020\|EPI_ISL_572851\|20 | ···················· | ···················· | ···················· | ···················· | ···················· | ···················· | |
| L | 1108 hCoV-19/England/CAMB-1B61F6/2020\|EPI_ISL_572852\|20 | ···················· | ···················· | ···················· | ···················· | ···················· | ···················· | |
| L | 1109 hCoV-19/Wales/PHWC-16C70C/2020\|EPI_ISL_572853\|2020 | ···················· | ···················· | ···················· | ···················· | ···················· | ···················· | |
| L | 1111 hCoV-19/Wales/PHWC-16C9E8/2020\|EPI_ISL_572855\|2020 | ···················· | ···················· | ···················· | ···················· | ···················· | ···················· | |
| L | 1112 hCoV-19/Wales/PHWC-16CB2E/2020\|EPI_ISL_572856\|2020 | ···················· | ···················· | ···················· | ···················· | ···················· | ···················· | |
| L | 1113 hCoV-19/England/ALDP-9B5DEC/2020\|EPI_ISL_572857\|20 | ···················· | ···················· | ···················· | ···················· | ···················· | ···················· | |
| L | 1114 hCoV-19/England/ALDP-9B5E52/2020\|EPI_ISL_572858\|20 | ···················· | ···················· | ···················· | ···················· | ···················· | ···················· | |
| L | 1115 hCoV-19/England/ALDP-9B60F1/2020\|EPI_ISL_572859\|20 | ···················· | ···················· | ···················· | ···················· | ···················· | ···················· | |
| L | 1116 hCoV-19/England/ALDP-9BCEC4/2020\|EPI_ISL_572860\|20 | ···················· | ···················· | ···················· | ···················· | ···················· | ···················· | |
| L | 1117 hCoV-19/England/ALDP-9BCEE2/2020\|EPI_ISL_572861\|20 | ···················· | ···················· | ···················· | ···················· | ···················· | ···················· | |
| L | 1119 hCoV-19/England/CAMB-1B5FA2/2020\|EPI_ISL_572863\|20 | ···················· | ···················· | ···················· | ···················· | ···················· | ···················· | |
| L | 1120 hCoV-19/England/CAMB-1B5FB1/2020\|EPI_ISL_572864\|20 | ···················· | ···················· | ···················· | ···················· | ···················· | ···················· | |
| L | 1121 hCoV-19/England/CAMB-1B5FC0/2020\|EPI_ISL_572865\|20 | ···················· | ···················· | ···················· | ···················· | ···················· | ···················· | |
| L | 1122 hCoV-19/England/CAMB-1B5FEE/2020\|EPI_ISL_572866\|20 | ···················· | ···················· | ···················· | ···················· | ···················· | ···················· | |
| L | 1123 hCoV-19/England/CAMB-1B619C/2020\|EPI_ISL_572867\|20 | ···················· | ···················· | ···················· | ···················· | ···················· | ···················· | |
| L | 1124 hCoV-19/England/CAMB-1B6211/2020\|EPI_ISL_572868\|20 | ···················· | ···················· | ···················· | ···················· | ···················· | ···················· | |
| L | 1125 hCoV-19/England/CAMC-9C0328/2020\|EPI_ISL_572869\|20 | ···················· | ···················· | ···················· | ···················· | ···················· | ···················· | |
| L | 1126 hCoV-19/England/CAMC-9C1697/2020\|EPI_ISL_572870\|20 | ···················· | ···················· | ···················· | ···················· | ···················· | ···················· | |
| L | 1127 hCoV-19/England/MILK-9A7FAB/2020\|EPI_ISL_572871\|20 | ···················· | ···················· | ···················· | ···················· | ···················· | ···················· | |
| L | 1128 hCoV-19/England/NOTT-11376E/2020\|EPI_ISL_572872\|20 | ···················· | ···················· | ···················· | ···················· | ···················· | ···················· | |
| L | 1129 hCoV-19/England/QEUH-9BA39A/2020\|EPI_ISL_572873\|20 | ···················· | ···················· | ···················· | ···················· | ···················· | ···················· | |
| L | 1130 hCoV-19/Wales/MILK-9A7FC9/2020\|EPI_ISL_572874\|2020 | ···················· | ···················· | ···················· | ···················· | ···················· | ···················· | |
| L | 1131 hCoV-19/Wales/PHWC-16B5F4/2020\|EPI_ISL_572875\|2020 | ···················· | ···················· | ···················· | ···················· | ···················· | ···················· | |
| L | 1132 hCoV-19/Wales/PHWC-16B6F1/2020\|EPI_ISL_572876\|2020 | ···················· | ···················· | ···················· | ···················· | ···················· | ···················· | |
| L | 1133 hCoV-19/Wales/PHWC-16BDCF/2020\|EPI_ISL_572877\|2020 | ···················· | ···················· | ···················· | ···················· | ···················· | ···················· | |
| L | 1134 hCoV-19/Wales/PHWC-16C03E/2020\|EPI_ISL_572878\|2020 | ···················· | ···················· | ···················· | ···················· | ···················· | ···················· | |
| L | 1135 hCoV-19/Wales/PHWC-16C46F/2020\|EPI_ISL_572879\|2020 | ···················· | ···················· | ···················· | ···················· | ···················· | ···················· | |
| L | 1136 hCoV-19/Wales/PHWC-16C4D8/2020\|EPI_ISL_572880\|2020 | ···················· | ···················· | ···················· | ···················· | ···················· | ···················· | |
| L | 1138 hCoV-19/Wales/PHWC-16C54E/2020\|EPI_ISL_572882\|2020 | ···················· | ···················· | ···················· | ···················· | ···················· | ···················· | |
| L | 1139 hCoV-19/Wales/PHWC-16C696/2020\|EPI_ISL_572883\|2020 | ···················· | ···················· | ···················· | ···················· | NNN················· | ···················· | 1 |

| | | | | | | | | |
|---|---|---|---|---|---|---|---|---|
| L | 1140 | hCoV-19/Wales/PHWC-16C757/2020\|EPI_ISL_572884\|2020 | ···················· | ···················· | ···················· | ···················· | ···················· | ···················· |
| L | 1141 | hCoV-19/Wales/PHWC-16CB3D/2020\|EPI_ISL_572885\|2020 | ···················· | ···················· | ···················· | ···················· | ···················· | ···················· |
| L | 1142 | hCoV-19/Wales/PHWC-16CEDA/2020\|EPI_ISL_572886\|2020 | ···················· | ···················· | ···················· | ···················· | ···················· | ···················· |
| L | 1143 | hCoV-19/Wales/PHWC-16D06A/2020\|EPI_ISL_572887\|2020 | ···················· | ···················· | ···················· | ···················· | ···················· | ···················· |
| L | 1145 | hCoV-19/Wales/PHWC-16D325/2020\|EPI_ISL_572889\|2020 | ···················· | ···················· | ···················· | ···················· | ···················· | ···················· |
| L | 1146 | hCoV-19/Wales/PHWC-16D334/2020\|EPI_ISL_572890\|2020 | ···················· | ···················· | ···················· | ···················· | ···················· | ···················· |
| L | 1147 | hCoV-19/USA/WA-SFS-S2852/2020\|EPI_ISL_575030\|2020- | ···················· | ·A·················· | ···················· | ···················· | ···················· | ···················· |
| L | 1148 | hCoV-19/USA/WA-SFS-S2853/2020\|EPI_ISL_575031\|2020- | ···················· | ·A·················· | ···················· | ···················· | ···················· | ···················· |
| L | 1149 | hCoV-19/USA/WA-SFS-S2854/2020\|EPI_ISL_575032\|2020- | ···················· | ·A··G··············· | ···················· | ···················· | ···················· | ···················· |
| L | 1150 | hCoV-19/USA/WA-SFS-S2855/2020\|EPI_ISL_575033\|2020- | ···················· | ···················· | ···················· | ···················· | ···················· | ···················· |
| L | 1151 | hCoV-19/USA/UT-UPHL-00017/2020\|EPI_ISL_575036\|2020 | ···················· | ···················· | ···················· | ···················· | ···················· | ···················· |
| L | 1152 | hCoV-19/USA/UN-UW-7196/2020\|EPI_ISL_576500\|2020-04 | ···················· | ···················· | ···················· | ···················· | ···················· | ···················· |
| L | 1153 | hCoV-19/India/MH-NIV-229643/2020\|EPI_ISL_577692\|20 | ···················· | ···················· | ···················· | ···················· | ···················· | ···················· |
| L | 1154 | hCoV-19/India/MH-NIV-229644/2020\|EPI_ISL_577693\|20 | ···················· | ···················· | ···················· | ···················· | ···················· | ···················· |
| L | 1155 | hCoV-19/India/MH-NIV-229715/2020\|EPI_ISL_577698\|20 | ···················· | ···················· | ···················· | ···················· | ···················· | ···················· |
| L | 1156 | hCoV-19/India/MH-NIV-229717/2020\|EPI_ISL_577699\|20 | ···················· | ···················· | ···················· | ···················· | ···················· | ···················· |
| L | 1157 | hCoV-19/mink/Netherlands/NB-EMC-44-5/2020\|EPI_ISL_ | ···················· | ···················· | ···················· | ···················· | ······T············· | ···················· |
| L | 1158 | hCoV-19/mink/Netherlands/NB-EMC-44-7/2020\|EPI_ISL_ | ···················· | ···················· | ···················· | ···················· | ······T············· | ···················· |
| L | 1159 | hCoV-19/mink/Netherlands/NB-EMC-45-3/2020\|EPI_ISL_ | ···················· | ···················· | ···················· | ···················· | ···················· | ···················· |
| L | 1160 | hCoV-19/mink/Netherlands/NB-EMC-45-4/2020\|EPI_ISL_ | ···················· | ···················· | ···················· | ···················· | ···················· | ···················· |
| L | 1161 | hCoV-19/mink/Netherlands/NB-EMC-46-10/2020\|EPI_ISL | ···················· | ···················· | ···················· | ···················· | ······T············· | ···················· |
| L | 1162 | hCoV-19/mink/Netherlands/NB-EMC-46-8/2020\|EPI_ISL_ | ···················· | ···················· | ···················· | ···················· | ······T············· | ···················· |
| L | 1163 | hCoV-19/mink/Netherlands/NB-EMC-47-1/2020\|EPI_ISL_ | ···················· | ···················· | ···················· | ···················· | ······T············· | ···················· |
| L | 1164 | hCoV-19/mink/Netherlands/NB-EMC-47-2/2020\|EPI_ISL_ | ···················· | ···················· | ···················· | ···················· | ······T············· | ···················· |
| L | 1165 | hCoV-19/mink/Netherlands/NB-EMC-47-5/2020\|EPI_ISL_ | ···················· | ···················· | ···················· | ···················· | ······T············· | ···················· |
| L | 1166 | hCoV-19/mink/Netherlands/NB-EMC-52-4/2020\|EPI_ISL_ | ···················· | ···················· | ···················· | ···················· | ······T············· | ···················· |
| L | 1167 | hCoV-19/Netherlands/GE-EMC-302/2020\|EPI_ISL_577829 | ···················· | ···················· | ···················· | ···················· | ······T············· | ···················· |
| L | 1168 | hCoV-19/Netherlands/GE-EMC-303/2020\|EPI_ISL_577830 | ···················· | ···················· | ···················· | ···················· | ···················· | ···················· |
| L | 1169 | hCoV-19/England/QEUH-9DC909/2020\|EPI_ISL_581011\|20 | ···················· | ·A·················· | ···················· | ···················· | ···················· | ··A················· |
| L | 1170 | hCoV-19/England/QEUH-9DCE28/2020\|EPI_ISL_581013\|20 | ···················· | ···················· | ···················· | ···················· | ···················· | ···················· |
| L | 1171 | hCoV-19/England/QEUH-9DC839/2020\|EPI_ISL_581014\|20 | ···················· | ···················· | ···················· | ···················· | ···················· | ···················· |
| L | 1172 | hCoV-19/England/QEUH-9DADC3/2020\|EPI_ISL_581015\|20 | ···················· | ···················· | ···················· | ···················· | ···················· | ···················· |
| L | 1173 | hCoV-19/England/QEUH-9DC8DF/2020\|EPI_ISL_581016\|20 | ···················· | ···················· | ···················· | ···················· | ···················· | ···················· |
| L | 1174 | hCoV-19/England/QEUH-9DC6E4/2020\|EPI_ISL_581017\|20 | ···················· | ···················· | ···················· | ···················· | ···················· | ···················· |
| L | 1175 | hCoV-19/England/MILK-9E0F97/2020\|EPI_ISL_581018\|20 | ···················· | ···················· | ···················· | ···················· | ···················· | ···················· |
| L | 1176 | hCoV-19/England/QEUH-9DC945/2020\|EPI_ISL_581019\|20 | ···················· | ·A·················· | ···················· | ···················· | ···················· | ··A················· |
| L | 1177 | hCoV-19/England/QEUH-9DAEC0/2020\|EPI_ISL_581020\|20 | ···················· | ···················· | ···················· | ···················· | ···················· | ···················· |
| L | 1178 | hCoV-19/England/QEUH-9DB0BA/2020\|EPI_ISL_581021\|20 | ···················· | ···················· | ···················· | ···················· | ···················· | ···················· |
| L | 1179 | hCoV-19/England/QEUH-9DC6C6/2020\|EPI_ISL_581022\|20 | ···················· | ···················· | ···················· | ···················· | ···················· | ···················· |
| L | 1180 | hCoV-19/England/MILK-9E10B1/2020\|EPI_ISL_581023\|20 | ···················· | ···················· | ···················· | ···················· | ···················· | ···················· |
| L | 1181 | hCoV-19/England/QEUH-9DC769/2020\|EPI_ISL_581024\|20 | ···················· | ···················· | ···················· | ···················· | ···················· | ···················· |
| L | 1182 | hCoV-19/England/MILK-9E0F79/2020\|EPI_ISL_581025\|20 | ···················· | ·A·················· | ···················· | ···················· | ···················· | ···················· |
| L | 1183 | hCoV-19/England/MILK-9E137C/2020\|EPI_ISL_581026\|20 | ···················· | ·A·················· | ···················· | ···················· | ···················· | ···················· |
| L | 1184 | hCoV-19/England/QEUH-9DCC79/2020\|EPI_ISL_581027\|20 | ···················· | ···················· | ···················· | ···················· | ···················· | ···················· |
| L | 1185 | hCoV-19/England/MILK-9E1075/2020\|EPI_ISL_581028\|20 | ···················· | ···················· | ···················· | ···················· | ···················· | ···················· |
| L | 1186 | hCoV-19/Wales/MILK-9E0FC4/2020\|EPI_ISL_581029\|2020 | ···················· | ···················· | ···················· | ···················· | ···················· | ···················· |
| L | 1187 | hCoV-19/England/QEUH-9DAC5D/2020\|EPI_ISL_581030\|20 | ···················· | ···················· | ···················· | ···················· | ···················· | ···················· |
| L | 1188 | hCoV-19/England/QEUH-9DC3B0/2020\|EPI_ISL_581031\|20 | ···················· | ···················· | ···················· | ···················· | ···················· | ···················· |
| L | 1189 | hCoV-19/England/MILK-9E0D51/2020\|EPI_ISL_581032\|20 | ···················· | ·A·················· | ···················· | ···················· | ···················· | ··A················· |
| L | 1190 | hCoV-19/England/QEUH-9DC936/2020\|EPI_ISL_581044\|20 | ···················· | ···················· | ···················· | ···················· | ···················· | ···················· |
| L | 1191 | hCoV-19/Wales/MILK-9E0F2E/2020\|EPI_ISL_581046\|2020 | ···················· | ···················· | ···················· | ···················· | ···················· | ···················· |
| L | 1192 | hCoV-19/England/QEUH-9DCB8B/2020\|EPI_ISL_581047\|20 | ···················· | ·A·················· | ···················· | ···················· | ···················· | ··A················· |
| L | 1193 | hCoV-19/England/QEUH-9DCFDA/2020\|EPI_ISL_581050\|20 | ···················· | ···················· | ···················· | ···················· | ···················· | ···················· |
| L | 1194 | hCoV-19/England/QEUH-9DC74B/2020\|EPI_ISL_581051\|20 | ···················· | ···················· | ···················· | ···················· | ···················· | ···················· |
| L | 1195 | hCoV-19/England/QEUH-9DC6F3/2020\|EPI_ISL_581052\|20 | ···················· | ···················· | ···················· | ···················· | ···················· | ···················· |
| L | 1196 | hCoV-19/England/QEUH-9DCA06/2020\|EPI_ISL_581053\|20 | ···················· | ···················· | ···················· | ···················· | ···················· | ···················· |
| L | 1197 | hCoV-19/Wales/MILK-9E1215/2020\|EPI_ISL_581054\|2020 | ···················· | ···················· | ···················· | ···················· | ···················· | ···················· |
| L | 1198 | hCoV-19/England/QEUH-9DAC4E/2020\|EPI_ISL_581055\|20 | ···················· | ···················· | ···················· | ···················· | ···················· | ···················· |
| L | 1199 | hCoV-19/England/MILK-9E12CA/2020\|EPI_ISL_581056\|20 | ···················· | ···················· | ···················· | ···················· | ···················· | ···················· |
| L | 1200 | hCoV-19/England/QEUH-9DC8A2/2020\|EPI_ISL_581057\|20 | ···················· | ·A·················· | ···················· | ···················· | ···················· | ··A················· |
| L | 1201 | hCoV-19/Wales/MILK-9E0FA6/2020\|EPI_ISL_581058\|2020 | ···················· | ···················· | ···················· | ···················· | ···················· | ···················· |
| L | 1202 | hCoV-19/England/QEUH-9DC56F/2020\|EPI_ISL_581059\|20 | ···················· | ···················· | ···················· | ···················· | ···················· | ···················· |
| L | 1203 | hCoV-19/England/QEUH-9DCE0A/2020\|EPI_ISL_581060\|20 | ···················· | ···················· | ···················· | ···················· | ···················· | ···················· |
| L | 1204 | hCoV-19/England/QEUH-9DD2A3/2020\|EPI_ISL_581061\|20 | ···················· | ·A·················· | ···················· | ···················· | ···················· | ··A················· |
| L | 1205 | hCoV-19/England/QEUH-9DC8FD/2020\|EPI_ISL_581062\|20 | ···················· | ···················· | ···················· | ···················· | ···················· | ···················· |
| L | 1206 | hCoV-19/England/QEUH-9DB0C9/2020\|EPI_ISL_581063\|20 | ···················· | ···················· | ···················· | ···················· | ···················· | ···················· |
| L | 1207 | hCoV-19/Wales/MILK-9E101B/2020\|EPI_ISL_581064\|2020 | ···················· | ···················· | ···················· | ···················· | ···················· | ···················· |
| L | 1208 | hCoV-19/England/MILK-9E0EC7/2020\|EPI_ISL_581065\|20 | ···················· | ···················· | ···················· | ···················· | ···················· | ···················· |
| L | 1209 | hCoV-19/Wales/MILK-9E127F/2020\|EPI_ISL_581067\|2020 | ···················· | ···················· | ···················· | ···················· | ···················· | ···················· |
| L | 1210 | hCoV-19/England/QEUH-9DC31A/2020\|EPI_ISL_581068\|20 | ···················· | ···················· | ···················· | ···················· | ···················· | ···················· |
| L | 1211 | hCoV-19/England/MILK-9E0ED6/2020\|EPI_ISL_581069\|20 | ···················· | ···················· | ···················· | ···················· | ······T············· | ···················· |
| L | 1212 | hCoV-19/England/QEUH-9DC66C/2020\|EPI_ISL_581070\|20 | ···················· | ···················· | ···················· | ···················· | ···················· | ···················· |
| L | 1213 | hCoV-19/England/QEUH-9DC72D/2020\|EPI_ISL_581071\|20 | ···················· | ···················· | ···················· | ···················· | ···················· | ···················· |
| L | 1214 | hCoV-19/England/QEUH-9DCE37/2020\|EPI_ISL_581072\|20 | ···················· | ···················· | ···················· | ···················· | ···················· | ···················· |
| L | 1215 | hCoV-19/England/QEUH-9DC893/2020\|EPI_ISL_581073\|20 | ···················· | ···················· | ···················· | ···················· | ···················· | ···················· |
| L | 1216 | hCoV-19/Wales/MILK-9E10FD/2020\|EPI_ISL_581074\|2020 | ···················· | ···················· | ···················· | ···················· | ···················· | ···················· |
| L | 1217 | hCoV-19/England/QEUH-9DADB4/2020\|EPI_ISL_581075\|20 | ···················· | ·A·················· | ···················· | ···················· | ···················· | ···················· |
| L | 1218 | hCoV-19/England/QEUH-9DABE7/2020\|EPI_ISL_581077\|20 | ···················· | ···················· | ···················· | ···················· | ···················· | ···················· |
| L | 1219 | hCoV-19/Wales/MILK-9E1172/2020\|EPI_ISL_581078\|2020 | ···················· | ···················· | ···················· | ···················· | ···················· | ···················· |
| L | 1220 | hCoV-19/England/QEUH-9DAE84/2020\|EPI_ISL_581079\|20 | ···················· | ···················· | ···················· | ···················· | ···················· | ···················· |

L 1221 hCoV-19/England/QEUH-9DC5AB/2020|EPI_ISL_581080|20 .................... .................... .................... .................... .................... ....................
L 1222 hCoV-19/England/QEUH-9DADE1/2020|EPI_ISL_581081|20 .................... .................... .................... .................... .................... ....................
L 1223 hCoV-19/England/QEUH-9DCB9A/2020|EPI_ISL_581082|20 .................... .................... .................... .................... .................... ....................
L 1224 hCoV-19/England/QEUH-9DC981/2020|EPI_ISL_581083|20 .................... .................... .................... .................... .................... ....................
L 1225 hCoV-19/Wales/MILK-9E1251/2020|EPI_ISL_581084|2020 .................... .................... .................... .................... .................... ....................
L 1226 hCoV-19/England/QEUH-9DCE82/2020|EPI_ISL_581085|20 .................... .................... .................... .................... .................... ....................
L 1227 hCoV-19/Wales/MILK-9E1206/2020|EPI_ISL_581086|2020 .................... .................... .................... .................... .................... ....................
L 1228 hCoV-19/England/QEUH-9DC57E/2020|EPI_ISL_581087|20 .................... .................... .................... .................... .................... ....................
L 1229 hCoV-19/England/QEUH-9DCA9D/2020|EPI_ISL_581089|20 .................... .A.................. .................... .................... .................... ...A................
L 1230 hCoV-19/England/QEUH-9DABD8/2020|EPI_ISL_581090|20 .................... .................... .................... .................... .................... ....................
L 1231 hCoV-19/England/QEUH-9DC9BE/2020|EPI_ISL_581091|20 .................... .................... .................... .................... .................... ....................
L 1232 hCoV-19/Wales/MILK-9E102A/2020|EPI_ISL_581093|2020 .................... .................... .................... .................... .................... ....................
L 1233 hCoV-19/England/QEUH-9DAD0F/2020|EPI_ISL_581094|20 .................... .................... .................... .................... .................... ....................
L 1234 hCoV-19/England/QEUH-9DAADB/2020|EPI_ISL_581095|20 .................... .................... .................... .................... .................... ....................
L 1235 hCoV-19/England/QEUH-9DAFDC/2020|EPI_ISL_581096|20 .................... .................... .................... .................... .................... ....................
L 1236 hCoV-19/England/MILK-9E1048/2020|EPI_ISL_581097|20 .................... .................... .................... .................... .................... ....................
L 1237 hCoV-19/England/QEUH-9DD30A/2020|EPI_ISL_581098|20 .................... .................... .................... .................... .................... ....................
L 1238 hCoV-19/Canada/MB-NML-859/2020|EPI_ISL_582342|2020 .................... .................... .................... .................... .................... ....................
L 1239 hCoV-19/Canada/MB-NML-861/2020|EPI_ISL_582343|2020 .................... .................... .................... .................... .................... ....................
L 1240 hCoV-19/Canada/MB-NML-864/2020|EPI_ISL_582345|2020 .................... .................... .................... .................... .................... ....................
L 1241 hCoV-19/Canada/MB-NML-865/2020|EPI_ISL_582346|2020 .................... .................... .................... .................... .................... ....................
L 1242 hCoV-19/Canada/MB-NML-866/2020|EPI_ISL_582347|2020 .................... .................... .................... .................... .................... ....................
L 1243 hCoV-19/Canada/MB-NML-872/2020|EPI_ISL_582349|2020 .................... .................... .................... .................... .................... ....................
L 1244 hCoV-19/Canada/MB-NML-873/2020|EPI_ISL_582350|2020 .................... .................... .................... .................... .................... ....................
L 1245 hCoV-19/Canada/MB-NML-874/2020|EPI_ISL_582351|2020 .................... .................... .................... .................... .................... ....................
L 1246 hCoV-19/Canada/MB-NML-876/2020|EPI_ISL_582352|2020 .................... .................... .................... .................... .................... ....................
L 1247 hCoV-19/Canada/MB-NML-877/2020|EPI_ISL_582353|2020 .................... .................... .................... .................... .................... ....................
L 1248 hCoV-19/Canada/MB-NML-878/2020|EPI_ISL_582354|2020 .................... .................... .................... .................... .................... ....................
L 1249 hCoV-19/Canada/MB-NML-880/2020|EPI_ISL_582355|2020 .................... .................... .................... .................... .................... ....................
L 1250 hCoV-19/Canada/MB-NML-979/2020|EPI_ISL_582356|2020 .................... .................... .................... .................... .................... ....................
L 1251 hCoV-19/Canada/MB-NML-980/2020|EPI_ISL_582357|2020 .................... .................... .................... .................... .................... ....................
L 1252 hCoV-19/Canada/MB-NML-981/2020|EPI_ISL_582358|2020 ...........T........ .................... .................... .................... .................... ....................
L 1253 hCoV-19/Canada/MB-NML-983/2020|EPI_ISL_582359|2020 .................... .A.................. .................... .................... .................... ...A................
L 1254 hCoV-19/Canada/MB-NML-984/2020|EPI_ISL_582360|2020 .................... .A.................. .................... .................... .................... ...A................
L 1255 hCoV-19/Canada/MB-NML-985/2020|EPI_ISL_582361|2020 .................... .A.................. .................... .................... .................... ...A................
L 1256 hCoV-19/Canada/MB-NML-987/2020|EPI_ISL_582362|2020 .................... .................... .................... .................... .................... ....................
L 1257 hCoV-19/Canada/MB-NML-989/2020|EPI_ISL_582363|2020 .................... .................... .................... .................... .................... ....................
L 1258 hCoV-19/Canada/MB-NML-1008/2020|EPI_ISL_582375|202 .................... .................... .................... .................... .................... ....................
L 1259 hCoV-19/Canada/MB-NML-1009/2020|EPI_ISL_582376|202 .................... .................... .................... .................... .................... ....................
L 1260 hCoV-19/Canada/MB-NML-1010/2020|EPI_ISL_582377|202 .................... .................... .................... .................... .................... ....................
L 1261 hCoV-19/Canada/MB-NML-1011/2020|EPI_ISL_582378|202 .................... .A.................. .................... .................... .................... ...A................
L 1262 hCoV-19/Canada/MB-NML-1014/2020|EPI_ISL_582380|202 .................... .................... .................... .................... .................... ....................
L 1263 hCoV-19/Canada/MB-NML-1015/2020|EPI_ISL_582381|202 .................... .................... .................... .................... .................... ....................
L 1264 hCoV-19/Canada/MB-NML-1016/2020|EPI_ISL_582382|202 .................... .................... .................... .................... .................... ....................
L 1265 hCoV-19/Canada/MB-NML-1017/2020|EPI_ISL_582383|202 .................... .................... .................... .................... .................... ....................
L 1266 hCoV-19/Canada/MB-NML-1018/2020|EPI_ISL_582384|202 .................... .................... .................... .................... .................... ....................
L 1267 hCoV-19/Canada/MB-NML-1019/2020|EPI_ISL_582385|202 .................... .................... .................... .................... .................... ....................
L 1268 hCoV-19/Canada/MB-NML-1020/2020|EPI_ISL_582386|202 .................... .A.................. .................... .................... .................... ...A................
L 1269 hCoV-19/Canada/MB-NML-1021/2020|EPI_ISL_582387|202 .................... .................... .................... .................... .................... ....................
L 1270 hCoV-19/Canada/MB-NML-1023/2020|EPI_ISL_582388|202 .................... .................... .................... .................... .................... ....................
L 1271 hCoV-19/Canada/MB-NML-1024/2020|EPI_ISL_582389|202 .................... .................... .................... .................... .................... ....................
L 1272 hCoV-19/Canada/MB-NML-1025/2020|EPI_ISL_582390|202 .................... .................... .................... .................... .................... ....................
L 1273 hCoV-19/Canada/MB-NML-1026/2020|EPI_ISL_582391|202 .................... .................... .................... .................... .................... ....................
L 1274 hCoV-19/Canada/MB-NML-1027/2020|EPI_ISL_582392|202 .................... .................... .................... .................... .................... ....................
L 1275 hCoV-19/Canada/MB-NML-1029/2020|EPI_ISL_582393|202 .................... .................... .................... .................... .................... ....................
L 1276 hCoV-19/Canada/MB-NML-1030/2020|EPI_ISL_582394|202 .................... .................... .................... .................... .................... ....................
L 1277 hCoV-19/Canada/MB-NML-1033/2020|EPI_ISL_582395|202 .................... .................... .................... .................... .................... ....................
L 1278 hCoV-19/Canada/MB-NML-1034/2020|EPI_ISL_582396|202 .................... .................... .................... .................... .................... ....................
L 1279 hCoV-19/Canada/MB-NML-1035/2020|EPI_ISL_582397|202 .................... .................... .................... .................... .................... ....................
L 1280 hCoV-19/Canada/MB-NML-1038/2020|EPI_ISL_582399|202 .................... .................... .................... .................... .................... ....................
L 1281 hCoV-19/USA/CA-QDX-874/2020|EPI_ISL_571573|2020-03 .................... .................... .................... .................... .................... ....................
L 1282 hCoV-19/USA/CA-QDX-915/2020|EPI_ISL_571574|2020-03 .................... .................... .................... .................... .................... ....................
L 1283 hCoV-19/USA/CA-QDX-916/2020|EPI_ISL_571575|2020-03 .................... .................... .................... .................... .................... ....................
L 1284 hCoV-19/USA/CA-QDX-920/2020|EPI_ISL_571576|2020-03 .................... .................... .................... .................... .................... ....................
L 1285 hCoV-19/USA/CA-QDX-921/2020|EPI_ISL_571577|2020-03 .................... .................... .................... .................... .................... ....................
L 1286 hCoV-19/USA/CA-QDX-922/2020|EPI_ISL_571578|2020-03 .................... .................... .................... .................... .................... ....................
L 1287 hCoV-19/USA/CA-QDX-924/2020|EPI_ISL_571580|2020-03 .................... .................... .................... .................... .................... ....................
L 1288 hCoV-19/USA/CA-QDX-929/2020|EPI_ISL_571581|2020-03 .................... .................... .................... .................... .................... ....................
L 1289 hCoV-19/USA/CA-QDX-936/2020|EPI_ISL_571582|2020-03 .................... .................... .................... .................... .................... ....................
L 1290 hCoV-19/USA/CA-QDX-938/2020|EPI_ISL_571583|2020-03 .................... .................... .................... .................... .................... ....................
L 1291 hCoV-19/USA/CA-QDX-939/2020|EPI_ISL_571584|2020-03 .................... .................... .................... .................... .................... ....................
L 1292 hCoV-19/USA/CA-QDX-940/2020|EPI_ISL_571585|2020-03 .................... .................... .................... .................... .................... ....................
L 1293 hCoV-19/USA/CA-QDX-941/2020|EPI_ISL_571586|2020-03 .................... .................... .................... .................... .................... ....................
L 1294 hCoV-19/USA/CA-QDX-942/2020|EPI_ISL_571587|2020-03 .................... .................... .................... .................... .................... ....................
L 1295 hCoV-19/USA/CA-QDX-944/2020|EPI_ISL_571588|2020-03 .................... .................... .................... .................... .................... ....................
L 1296 hCoV-19/USA/CA-QDX-945/2020|EPI_ISL_571589|2020-03 .................... .................... .................... .................... .................... ....................
L 1297 hCoV-19/USA/CA-QDX-946/2020|EPI_ISL_571590|2020-03 .................... .................... .................... .................... .................... ....................
L 1298 hCoV-19/USA/CA-QDX-948/2020|EPI_ISL_571591|2020-03 .................... .................... .................... .................... .................... ....................
L 1299 hCoV-19/USA/CO-QDX-877/2020|EPI_ISL_571592|2020-03 .................... .................... .................... .................... .................... ....................
L 1300 hCoV-19/USA/FL-QDX-909/2020|EPI_ISL_571593|2020-03 .................... .................... .................... .................... .................... ....................

L 1301 hCoV-19/USA/FL-QDX-910/2020|EPI_ISL_571594|2020-03 ···················· ···················· ···················· ···················· ···················· ····················
L 1302 hCoV-19/Switzerland/ZH-ETHZ-100214/2020|EPI_ISL_57 ···················· ···················· ···················· ···················· ···················· W···················
L 1303 hCoV-19/Switzerland/ZH-ETHZ-100242/2020|EPI_ISL_57 ···················· ···················· ···················· ···················· ···················· ····················
L 1304 hCoV-19/Switzerland/ZH-ETHZ-100243/2020|EPI_ISL_57 ···················· ···················A ···················· ···················· ···················· ···················· 1
L 1305 hCoV-19/Switzerland/ZH-ETHZ-100370/2020|EPI_ISL_57 ···················· ···················· ···················· ···················· ···················· ····················
L 1306 hCoV-19/USA/WA-SFS-S2850/2020|EPI_ISL_575028|2020- ···················· ···················· ···················· ···················· ···················· ····················
L 1307 hCoV-19/USA/WA-SFS-S2851/2020|EPI_ISL_575029|2020- ···················· ·A·················· ···················· ···················· ···················· ····················
L 1308 hCoV-19/USA/UT-UPHL-00015/2020|EPI_ISL_575035|2020 ···················· ···················· ···················· ···················· ···················· ····················
L 1309 hCoV-19/USA/UT-UPHL-01354/2020|EPI_ISL_575038|2020 ···················· ···················· ···················· ···················· ···················· ····················
L 1310 hCoV-19/USA/UT-UPHL-01358/2020|EPI_ISL_575039|2020 ···················· ···················· ···················· ···················· ···················· ····················
L 1311 hCoV-19/USA/UT-UPHL-01361/2020|EPI_ISL_575040|2020 ···················· ···················· ···················· ···················· ···················· ····················
L 1312 hCoV-19/USA/UT-UPHL-01363/2020|EPI_ISL_575041|2020 ···················· ···················· ···················· ···················· ···················· ····················
L 1313 hCoV-19/USA/UT-UPHL-01375/2020|EPI_ISL_575043|2020 ···················· ···················· ···················· ···················· ···················· ····················
L 1314 hCoV-19/USA/UT-UPHL-01379/2020|EPI_ISL_575044|2020 ···················· ···················· ···················· ···················· ···················· ····················
L 1315 hCoV-19/USA/UT-UPHL-01382/2020|EPI_ISL_575045|2020 ···················· ···················· ···················· ···················· ···················· ····················
L 1316 hCoV-19/USA/UT-UPHL-01384/2020|EPI_ISL_575046|2020 ···················· ···················· ···················· ···················· ···················· ····················
L 1317 hCoV-19/USA/UT-UPHL-01388/2020|EPI_ISL_575047|2020 ···················· ···················· ···················· ···················· ···················· ····················
L 1318 hCoV-19/USA/UT-UPHL-01393/2020|EPI_ISL_575048|2020 ···················· ···················· ···················· ···················· ···················· ····················
L 1319 hCoV-19/USA/UT-UPHL-01396/2020|EPI_ISL_575049|2020 ···················· ···················· ···················· ···················· ···················· ····················
L 1320 hCoV-19/USA/UT-UPHL-01398/2020|EPI_ISL_575050|2020 ···················· ···················· ···················· ···················· ···················· ····················
L 1321 hCoV-19/USA/UT-UPHL-01402/2020|EPI_ISL_575052|2020 ···················· ···················· ···················· ···················· ···················· ····················
L 1322 hCoV-19/USA/UT-UPHL-01405/2020|EPI_ISL_575053|2020 ···················· ···················· ···················· ···················· ···················· ····················
L 1323 hCoV-19/USA/UT-UPHL-01407/2020|EPI_ISL_575054|2020 ···················· ···················· ···················· ···················· ···················· ····················
L 1324 hCoV-19/USA/UT-UPHL-01408/2020|EPI_ISL_575055|2020 ···················· ···················· ···················· ···················· ···················· ····················
L 1325 hCoV-19/USA/UT-UPHL-01415/2020|EPI_ISL_575057|2020 ···················· ···················· ···················· ···················· ···················· ····················
L 1326 hCoV-19/USA/UT-UPHL-01419/2020|EPI_ISL_575058|2020 ···················· ···················· ···················· ···················· ···················· ····················
L 1327 hCoV-19/USA/UT-UPHL-01421/2020|EPI_ISL_575059|2020 ···················· ···················· ···················· ···················· ···················· ····················
L 1328 hCoV-19/USA/UT-UPHL-01423/2020|EPI_ISL_575060|2020 ···················· ···················· ···················· ···················· ···················· ····················
L 1329 hCoV-19/USA/UT-UPHL-01635/2020|EPI_ISL_575061|2020 ···················· ···················· ···················· ···················· ···················· ····················
L 1330 hCoV-19/USA/UT-UPHL-01637/2020|EPI_ISL_575063|2020 ···················· ···················· ···················· ···················· ···················· ····················
L 1331 hCoV-19/USA/UT-UPHL-01640/2020|EPI_ISL_575065|2020 ···················· ···················· ···················· ···················· ···················· ····················
L 1332 hCoV-19/USA/UT-UPHL-01643/2020|EPI_ISL_575067|2020 ···················· ···················· ···················· ···················· ···················· ····················
L 1333 hCoV-19/USA/UT-UPHL-01644/2020|EPI_ISL_575068|2020 ···················· ···················· ···················· ···················· ···················· ····················
L 1334 hCoV-19/USA/UT-UPHL-01645/2020|EPI_ISL_575069|2020 ···················· ···················· ···················· ···················· ···················· ····················
L 1335 hCoV-19/USA/UT-UPHL-02763/2020|EPI_ISL_575075|2020 ···················· ···················· ···················· ···················· ···················· ····················
L 1336 hCoV-19/USA/UT-UPHL-02802/2020|EPI_ISL_575079|2020 ···················· ···················· ···················· ···················· ···················· ····················
L 1337 hCoV-19/USA/UT-UPHL-02805/2020|EPI_ISL_575080|2020 ···················· ···················· ···················· ···················· ···················· ····················
L 1338 hCoV-19/USA/UT-UPHL-02807/2020|EPI_ISL_575081|2020 ···················· ···················· ···················· ···················· ···················· ····················
L 1339 hCoV-19/USA/UT-UPHL-02817/2020|EPI_ISL_575083|2020 ···················· ···················· ···················· ···················· ···················· ····················
L 1340 hCoV-19/USA/UT-UPHL-02819/2020|EPI_ISL_575084|2020 ···················· ···················· ···················· ···················· ···················· ····················
L 1341 hCoV-19/USA/UT-UPHL-02822/2020|EPI_ISL_575085|2020 ···················· ···················· ···················· ···················· ···················· ····················
L 1342 hCoV-19/USA/UT-UPHL-02824/2020|EPI_ISL_575086|2020 ···················· ···················· ···················· ···················· ···················· ····················
L 1343 hCoV-19/USA/UT-UPHL-02827/2020|EPI_ISL_575087|2020 ···················· ···················· ···················· ···················· ···················· ····················
L 1344 hCoV-19/USA/UT-UPHL-02828/2020|EPI_ISL_575088|2020 ···················· ···················· ···················· ···················· ···················· ····················
L 1345 hCoV-19/USA/UT-UPHL-02829/2020|EPI_ISL_575089|2020 ···················· ···················· ···················· ···················· ···················· ····················
L 1346 hCoV-19/USA/UT-UPHL-02830/2020|EPI_ISL_575090|2020 ···················· ···················· ···················· ···················· ···················· ····················
L 1347 hCoV-19/USA/UT-UPHL-02831/2020|EPI_ISL_575091|2020 ···················· ···················· ···················· ···················· ···················· ····················
L 1348 hCoV-19/USA/UT-UPHL-02832/2020|EPI_ISL_575092|2020 ···················· ···················· ···················· ···················· ···················· ····················
L 1349 hCoV-19/USA/UT-UPHL-02833/2020|EPI_ISL_575093|2020 ···················· ···················· ···················· ···················· ···················· ····················
L 1350 hCoV-19/USA/UT-UPHL-03746/2020|EPI_ISL_575099|2020 ···················· ···················· ···················· ···················· ···················· ····················
L 1351 hCoV-19/USA/UT-UPHL-03749/2020|EPI_ISL_575100|2020 ···················· ···················· ···················· ···················· ···················· ····················
L 1352 hCoV-19/USA/UT-UPHL-03750/2020|EPI_ISL_575101|2020 ···················· ···················· ···················· ···················· ···················· ····················
L 1353 hCoV-19/USA/UT-UPHL-03751/2020|EPI_ISL_575102|2020 ···················· ···················· ···················· ···················· ···················· ····················
L 1354 hCoV-19/USA/UT-UPHL-03753/2020|EPI_ISL_575103|2020 ···················· ···················· ···················· ···················· ···················· ····················
L 1355 hCoV-19/USA/UT-UPHL-03754/2020|EPI_ISL_575104|2020 ···················· ···················· ···················· ···················· ···················· ····················
L 1356 hCoV-19/USA/UT-UPHL-03756/2020|EPI_ISL_575106|2020 ···················· ···················· ···················· ···················· ···················· ····················
L 1357 hCoV-19/USA/UT-UPHL-03757/2020|EPI_ISL_575107|2020 ···················· ···················· ···················· ···················· ···················· ····················
L 1358 hCoV-19/USA/UT-UPHL-03758/2020|EPI_ISL_575108|2020 ···················· ···················· ···················· ···················· ···················· ····················
L 1359 hCoV-19/USA/UT-UPHL-03760/2020|EPI_ISL_575109|2020 ···················· ···················· ···················· ···················· ···················· ····················
L 1360 hCoV-19/USA/UT-UPHL-03763/2020|EPI_ISL_575111|2020 ···················· ···················· ···················· ···················· ···················· ····················
L 1361 hCoV-19/USA/UT-UPHL-03764/2020|EPI_ISL_575112|2020 ···················· ···················· ···················· ···················· ···················· ····················
L 1362 hCoV-19/USA/UT-UPHL-04230/2020|EPI_ISL_575119|2020 ···················· ···················· ···················· ···················· ···················· ····················
L 1363 hCoV-19/USA/UT-UPHL-04235/2020|EPI_ISL_575122|2020 ···················· ···················· ···················· ···················· ···················· ····················
L 1364 hCoV-19/USA/UT-UPHL-04236/2020|EPI_ISL_575123|2020 ···················· ···················· ···················· ···················· ···················· ····················
L 1365 hCoV-19/USA/UT-UPHL-04262/2020|EPI_ISL_575125|2020 ···················· ···················· ···················· ···················· ···················· ····················
L 1366 hCoV-19/USA/UT-UPHL-04265/2020|EPI_ISL_575126|2020 ···················· ···················· ···················· ···················· ···················· ····················
L 1367 hCoV-19/USA/UT-UPHL-04282/2020|EPI_ISL_575127|2020 ···················· ···················· ···················· ···················· ···················· ····················
L 1368 hCoV-19/USA/UT-UPHL-04289/2020|EPI_ISL_575130|2020 ···················· ···················· ···················· ···················· ···················· ····················
L 1369 hCoV-19/USA/UT-UPHL-04290/2020|EPI_ISL_575131|2020 ···················· ···················· ···················· ···················· ···················· ····················
L 1370 hCoV-19/USA/UT-UPHL-04306/2020|EPI_ISL_575136|2020 ···················· ···················· ···················· ···················· ···················· ····················
L 1371 hCoV-19/USA/TX-DSHS-0710/2020|EPI_ISL_576353|2020- ···················· ·A·················· ···················· ···················· ···················· ····················
L 1372 hCoV-19/Egypt/CUNCI-HGC7I010/2020|EPI_ISL_576371|2 ···················· ···················· ···················· ···················· ···················· ····················
L 1373 hCoV-19/Egypt/CUNCI-HGC7I023/2020|EPI_ISL_576372|2 ···················· ···················· ···················· ···················· ···················· ····················
L 1374 hCoV-19/Egypt/CUNCI-HGC7I025/2020|EPI_ISL_576373|2 ···················· ···················· ···················· ···················· ···················· ····················
L 1375 hCoV-19/Indonesia/JT-UGM-47906/2020|EPI_ISL_576383 ···················· ···················· ···················· ···················· ···················· ····················
L 1376 hCoV-19/Indonesia/JK-NIHRD-MI43269/2020|EPI_ISL_57 ···················· ···················· ···················· ···················· ···················· ····················
L 1377 hCoV-19/Indonesia/JK-NIHRD-MI52948/2020|EPI_ISL_57 ···················· ···················· ···················· ···················· ···················· ····················
L 1378 hCoV-19/USA/UN-UW-10411/2020|EPI_ISL_576399|2020-0 ···················· ···················· ···················· ···················· ···················· ····················
L 1379 hCoV-19/USA/UN-UW-11208/2020|EPI_ISL_576401|2020-0 ···················· ···················· ···················· ···················· ···················· ····················
L 1380 hCoV-19/USA/UN-UW-11929/2020|EPI_ISL_576402|2020-0 ···················· ···················· ···················· ···················· ···················· ····················

L 1381 hCoV-19/USA/UN-UW-12776/2020|EPI_ISL_576403|2020-0 ···················· ···················· ····················· ···················· ···················· ····················
L 1382 hCoV-19/USA/UN-UW-12782/2020|EPI_ISL_576404|2020-0 ···················· ···················· ····················· ···················· ···················· ····················
L 1383 hCoV-19/USA/WA-UW-12787/2020|EPI_ISL_576405|2020-0 ···················· ·A·················· ····················· ···················· ···················· ····················
L 1384 hCoV-19/USA/WA-UW-23234/2020|EPI_ISL_576408|2020-0 ···················· ·A·················· ····················· ···················· ···················· ····················
L 1385 hCoV-19/USA/WA-UW-23235/2020|EPI_ISL_576409|2020-0 ···················· ·A·················· ····················· ···················· ···················· ····················
L 1386 hCoV-19/USA/WA-UW-23237/2020|EPI_ISL_576410|2020-0 ···················· ···················· ····················· ···················· ···················· ····················
L 1387 hCoV-19/USA/WA-UW-23252/2020|EPI_ISL_576411|2020-0 ···················· ·A·················· ····················· ···················· ···················· ····················
L 1388 hCoV-19/USA/WA-UW-23257/2020|EPI_ISL_576412|2020-0 ···················· ···················· ····················· ···················· ···················· ····················
L 1389 hCoV-19/USA/WA-UW-23276/2020|EPI_ISL_576414|2020-0 ···················· ·A·················· ····················· ···················· ···················· ····················
L 1390 hCoV-19/USA/WA-UW-23277/2020|EPI_ISL_576415|2020-0 ···················· ···················· ····················· ···················· ···················· ····················
L 1391 hCoV-19/USA/WA-UW-23278/2020|EPI_ISL_576416|2020-0 ···················· ···················· ····················· ···················· ···················· ····················
L 1392 hCoV-19/USA/WA-UW-23283/2020|EPI_ISL_576417|2020-0 ···················· ···················· ····················· ···················· ···················· ····················
L 1393 hCoV-19/USA/WA-UW-23293/2020|EPI_ISL_576418|2020-0 ···················· ·A·················· ····················· ···················· ···················· ····················
L 1394 hCoV-19/USA/WA-UW-23297/2020|EPI_ISL_576419|2020-0 ···················· ·A·················· ····················· ···················· ···················· ····················
L 1395 hCoV-19/USA/WA-UW-23299/2020|EPI_ISL_576420|2020-0 ···················· ···················· ····················· ···················· ···················· ····················
L 1396 hCoV-19/USA/WA-UW-23305/2020|EPI_ISL_576422|2020-0 ···················· ···················· ····················· ···················· ···················· ····················
L 1397 hCoV-19/USA/WA-UW-23308/2020|EPI_ISL_576423|2020-0 ···················· ·A·················· ····················· ···················· ···················· ····················
L 1398 hCoV-19/USA/WA-UW-23312/2020|EPI_ISL_576424|2020-0 ···················· ···················· ····················· ···················· ···················· ····················
L 1399 hCoV-19/USA/WA-UW-23313/2020|EPI_ISL_576425|2020-0 ···················· ···················· ····················· ···················· ···················· ····················
L 1400 hCoV-19/USA/WA-UW-23330/2020|EPI_ISL_576426|2020-0 ···················· ···················· ····················· ···················· ···················· ····················
L 1401 hCoV-19/USA/OR-UW-23339/2020|EPI_ISL_576427|2020-0 ···················· ···················· ····················· ···················· ···················· ····················
L 1402 hCoV-19/USA/WA-UW-23340/2020|EPI_ISL_576428|2020-0 ···················· ·A·················· ····················· ···················· ···················· ····················
L 1403 hCoV-19/USA/WA-UW-23342/2020|EPI_ISL_576429|2020-0 ···················· ···················· ····················· ···················· ···················· ····················
L 1404 hCoV-19/USA/WA-UW-23349/2020|EPI_ISL_576430|2020-0 ···················· ···················· ····················· ···················· ···················· ····················
L 1405 hCoV-19/USA/WA-UW-23355/2020|EPI_ISL_576431|2020-0 ···················· ···················· ····················· ···················· ···················· ····················
L 1406 hCoV-19/USA/WA-UW-23360/2020|EPI_ISL_576432|2020-0 ···················· ···················· ····················· ···················· ···················· ····················
L 1407 hCoV-19/USA/WA-UW-23365/2020|EPI_ISL_576433|2020-0 ···················· ·A·················· ····················· ···················· ···················· ····················
L 1408 hCoV-19/USA/WA-UW-23371/2020|EPI_ISL_576434|2020-0 ···················· ·A·················· ····················· ···················· ···················· ····················
L 1409 hCoV-19/USA/WA-UW-23376/2020|EPI_ISL_576435|2020-0 ···················· ···················· ····················· ···················· ···················· ····················
L 1410 hCoV-19/USA/WA-UW-23378/2020|EPI_ISL_576436|2020-0 ···················· ···················· ····················· ···················· ···················· ····················
L 1411 hCoV-19/USA/WA-UW-23379/2020|EPI_ISL_576437|2020-0 ···················· ·A·················· ····················· ···················· ···················· ····················
L 1412 hCoV-19/USA/WA-UW-23380/2020|EPI_ISL_576438|2020-0 ···················· ···················· ····················· ···················· ···················· ····················
L 1413 hCoV-19/USA/WA-UW-23384/2020|EPI_ISL_576439|2020-0 ···················· ···················· ····················· ···················· ···················· ····················
L 1414 hCoV-19/USA/WA-UW-23389/2020|EPI_ISL_576440|2020-0 ···················· ···················· ····················· ···················· ···················· ····················
L 1415 hCoV-19/USA/WA-UW-23390/2020|EPI_ISL_576441|2020-0 ···················· ·A·················· ····················· ···················· ···················· ····················
L 1416 hCoV-19/USA/WA-UW-23391/2020|EPI_ISL_576442|2020-0 ···················· ···················· ····················· ···················· ···················· ····················
L 1417 hCoV-19/USA/WA-UW-23394/2020|EPI_ISL_576443|2020-0 ···················· ···················· ····················· ···················· ···················· ····················
L 1418 hCoV-19/USA/WA-UW-23397/2020|EPI_ISL_576444|2020-0 ···················· ·A·················· ····················· ···················· ···················· ····················
L 1419 hCoV-19/USA/WA-UW-23400/2020|EPI_ISL_576445|2020-0 ···················· ···················· ····················· ···················· ···················· ····················
L 1420 hCoV-19/USA/WA-UW-23406/2020|EPI_ISL_576446|2020-0 ···················· ···················· ····················· ···················· ···················· ····················
L 1421 hCoV-19/USA/WA-UW-23410/2020|EPI_ISL_576447|2020-0 ···················· ···················· ····················· ···················· ···················· ····················
L 1422 hCoV-19/USA/WA-UW-23414/2020|EPI_ISL_576448|2020-0 ···················· ···················· ····················· ···················· ···················· ····················
L 1423 hCoV-19/USA/WA-UW-23415/2020|EPI_ISL_576449|2020-0 ···················· ·A·················· ····················· ···················· ···················· ····················
L 1424 hCoV-19/USA/WA-UW-23422/2020|EPI_ISL_576450|2020-0 ···················· ·A·················· ····················· ···················· ···················· ····················
L 1425 hCoV-19/USA/WA-UW-23423/2020|EPI_ISL_576451|2020-0 ···················· ···················· ····················· ···················· ···················· ····················
L 1426 hCoV-19/USA/WA-UW-23429/2020|EPI_ISL_576452|2020-0 ···················· ·A·················· ····················· ···················· ···················· ····················
L 1427 hCoV-19/USA/WA-UW-23430/2020|EPI_ISL_576453|2020-0 ···················· ···················· ····················· ···················· ···················· ····················
L 1428 hCoV-19/USA/WA-UW-23431/2020|EPI_ISL_576454|2020-0 ···················· ···················· ····················· ···················· ···················· ····················
L 1429 hCoV-19/USA/WA-UW-23445/2020|EPI_ISL_576455|2020-0 ···················· ···················· ····················· ···················· ···················· ····················
L 1430 hCoV-19/USA/WA-UW-23461/2020|EPI_ISL_576456|2020-0 ···················· ·A·················· ····················· ···················· ···················· ····················
L 1431 hCoV-19/USA/WA-UW-23464/2020|EPI_ISL_576457|2020-0 ···················· ·A·················· ····················· ···················· ···················· ····················
L 1432 hCoV-19/USA/WA-UW-23467/2020|EPI_ISL_576458|2020-0 ···················· ···················· ····················· ···················· ···················· ····················
L 1433 hCoV-19/USA/WA-UW-23468/2020|EPI_ISL_576459|2020-0 ···················· ···················· ····················· ···················· ···················· ····················
L 1434 hCoV-19/USA/WA-UW-23476/2020|EPI_ISL_576460|2020-0 ···················· ···················· ····················· ···················· ···················· ····················
L 1435 hCoV-19/USA/WA-UW-23477/2020|EPI_ISL_576461|2020-0 ···················· ···················· ····················· ···················· ···················· ····················
L 1436 hCoV-19/USA/WA-UW-23481/2020|EPI_ISL_576462|2020-0 ···················· ·A·················· ····················· ···················· ···················· ····················
L 1437 hCoV-19/USA/WA-UW-23487/2020|EPI_ISL_576463|2020-0 ···················· ···················· ····················· ···················· ···················· ····················
L 1438 hCoV-19/USA/WA-UW-23488/2020|EPI_ISL_576464|2020-0 ···················· ·A·················· ····················· ···················· ···················· ····················
L 1439 hCoV-19/USA/WA-UW-23489/2020|EPI_ISL_576465|2020-0 ···················· ···················· ····················· ···················· ···················· ····················
L 1440 hCoV-19/USA/WA-UW-23498/2020|EPI_ISL_576466|2020-0 ···················· ···················· ····················· ···················· ···················· ····················
L 1441 hCoV-19/USA/WA-UW-23501/2020|EPI_ISL_576467|2020-0 ···················· ·A·················· ····················· ···················· ···················· ····················
L 1442 hCoV-19/India/MH-NIV-217242/2020|EPI_ISL_577678|20 ···················· ···················· ····················· ···················· ···················· ····················
L 1443 hCoV-19/India/MH-NIV-229467/2020|EPI_ISL_577679|20 ···················· ···················· ····················· ···················· ···················· ····················
L 1444 hCoV-19/India/MH-NIV-229468/2020|EPI_ISL_577680|20 ···················· ···················· ····················· ···················· ···················· ····················
L 1445 hCoV-19/India/MH-NIV-229461/2020|EPI_ISL_577681|20 ···················· ···················· ····················· ···················· ···················· ····················
L 1446 hCoV-19/India/MH-NIV-229428/2020|EPI_ISL_577682|20 ···················· ···················· ····················· ···················· ···················· ····················
L 1447 hCoV-19/India/MH-NIV-229512/2020|EPI_ISL_577683|20 ···················· ·A·················· ····················· ···················· ···················· ··A·················
L 1448 hCoV-19/India/MH-NIV-229539/2020|EPI_ISL_577684|20 ···················· ···················· ····················· ···················· ···················· ····················
L 1449 hCoV-19/India/MH-NIV-229542/2020|EPI_ISL_577685|20 ···················· ···················· ····················· ···················· ···················· ····················
L 1450 hCoV-19/India/MH-NIV-229545/2020|EPI_ISL_577686|20 ···················· ···················· ····················· ···················· ···················· ····················
L 1451 hCoV-19/India/MH-NIV-229566/2020|EPI_ISL_577688|20 ···················· ···················· ····················· ···················· ···················· ····················
L 1452 hCoV-19/India/MH-NIV-229576/2020|EPI_ISL_577689|20 ···················· ···················· ····················· ···················· ···················· ····················
L 1453 hCoV-19/India/MH-NIV-229597/2020|EPI_ISL_577690|20 ···················· ···················· ····················· ···················· ···················· ····················
L 1454 hCoV-19/India/MH-NIV-229622/2020|EPI_ISL_577691|20 ···················· ···················· ···········T········· ···················· ···················· ····················
L 1455 hCoV-19/India/MH-NIV-229665/2020|EPI_ISL_577695|20 ············T······· ···················· ····················· ···················· ···················· ····················
L 1456 hCoV-19/India/MH-NIV-229360/2020|EPI_ISL_577711|20 ···················· ···················· ····················· ···················· ···················· ····················
L 1457 hCoV-19/India/MH-NIV-229453/2020|EPI_ISL_577712|20 ···················· ···················· ····················· ···················· ···················· ····················
L 1458 hCoV-19/India/MH-NIV-229460/2020|EPI_ISL_577713|20 ···················· ···················· ····················· ···················· ···················· ····················
L 1459 hCoV-19/India/KA-NIV-QC-1803/2020|EPI_ISL_577714|2 ···················· ·A·················· ····················· ···················· ···················· ··A·················
L 1460 hCoV-19/India/KA-NIV-QC-1801/2020|EPI_ISL_577715|2 ···················· ·A·················· ····················· ···················· ···················· ··A·················

| | | | | | | | | |
|---|---|---|---|---|---|---|---|---|
| L | 1461 | hCoV-19/India/MH-NIV-229388/2020\|EPI_ISL_577717\|20 | ···················· | ···················· | ···················· | ···················· | ···················· | ···················· |
| L | 1462 | hCoV-19/India/MH-NIV-227726/2020\|EPI_ISL_577718\|20 | ···················· | ···················· | ···················· | ···················· | ···················· | ···················· |
| L | 1463 | hCoV-19/India/MH-NIV-226708/2020\|EPI_ISL_577719\|20 | ···················· | ···················· | ···················· | ···················· | ···················· | ···················· |
| L | 1464 | hCoV-19/India/MH-NIV-226709/2020\|EPI_ISL_577720\|20 | ···················· | ···················· | ···················· | ···················· | ···················· | ···················· |
| L | 1465 | hCoV-19/India/MH-NIV-227243/2020\|EPI_ISL_577722\|20 | ···················· | ·A·················· | ···················· | ···················· | ···················· | ··A················· |
| L | 1466 | hCoV-19/India/MH-NIV-227512/2020\|EPI_ISL_577723\|20 | ···················· | ···················· | ···················· | ···················· | ···················· | ···················· |
| L | 1467 | hCoV-19/India/MH-NIV-QC-1379/2020\|EPI_ISL_577725\|2 | ···················· | ···················· | ···················· | ···················· | ···················· | ···················· |
| L | 1468 | hCoV-19/India/MH-NIV-QC-1381/2020\|EPI_ISL_577726\|2 | ···················· | ···················· | ···················· | ···················· | ···················· | ···················· |
| L | 1469 | hCoV-19/India/MH-NIV-QC-1444/2020\|EPI_ISL_577727\|2 | ···················· | ·A·················· | ···················· | ···················· | ···················· | ··A················· |
| L | 1470 | hCoV-19/India/MH-NIV-QC-1450/2020\|EPI_ISL_577728\|2 | ···················· | ···················· | ···················· | ···················· | ···················· | ···················· |
| L | 1471 | hCoV-19/India/MH-NIV-QC-1405/2020\|EPI_ISL_577729\|2 | ···················· | ···················· | ···················· | ···················· | ···················· | ···················· |
| L | 1472 | hCoV-19/India/MH-NIV-QC-1844/2020\|EPI_ISL_577730\|2 | ···················· | ···················· | ···················· | ···················· | ···················· | ···················· |
| L | 1473 | hCoV-19/India/MH-NIV-QC-1848/2020\|EPI_ISL_577731\|2 | ···················· | ···················· | ···················· | ···················· | ···················· | ···················· |
| L | 1474 | hCoV-19/India/MH-NIV-QC-1822/2020\|EPI_ISL_577732\|2 | ···················· | ···················· | ···················· | ···················· | ···················· | ···················· |
| L | 1475 | hCoV-19/India/MH-NIV-QC-1804/2020\|EPI_ISL_577733\|2 | ···················· | ···················· | ···················· | ···················· | ···················· | ···················· |
| L | 1476 | hCoV-19/Slovakia/UKBA-201/2020\|EPI_ISL_577734\|2020 | ···················· | ···················· | ···················· | ···················· | ···················· | ···················· |
| L | 1477 | hCoV-19/Slovakia/UKBA-203/2020\|EPI_ISL_577735\|2020 | ···················· | ···················· | ···················· | ···················· | ···················· | ···················· |
| L | 1478 | hCoV-19/Slovakia/UKBA-204/2020\|EPI_ISL_577736\|2020 | ···················· | ···················· | ···················· | ···················· | ···················· | ···················· |
| L | 1479 | hCoV-19/Slovakia/UKBA-205/2020\|EPI_ISL_577737\|2020 | ···················· | ···················· | ···················· | ···················· | ···················· | ···················· |
| L | 1480 | hCoV-19/Slovakia/UKBA-207/2020\|EPI_ISL_577738\|2020 | ···················· | ···················· | ···················· | ···················· | ···················· | ···················· |
| L | 1481 | hCoV-19/Slovakia/UKBA-208/2020\|EPI_ISL_577739\|2020 | ···················· | ···················· | ···················· | ···················· | ···················· | ···················· |
| L | 1482 | hCoV-19/Slovakia/UKBA-209/2020\|EPI_ISL_577740\|2020 | ···················· | ···················· | ···················· | ···················· | ···················· | ···················· |
| L | 1483 | hCoV-19/Slovakia/UKBA-210/2020\|EPI_ISL_577741\|2020 | ···················· | ···················· | ···················· | ···················· | ···················· | ···················· |
| L | 1484 | hCoV-19/Slovakia/UKBA-212/2021\|EPI_ISL_577742\|2020 | ···················· | ···················· | ···················· | ···················· | ···················· | ···················· |
| L | 1485 | hCoV-19/Belgium/un-EMC-1/2020\|EPI_ISL_577743\|2020- | ···················· | ···················· | ···················· | ···················· | ···················· | ···················· |
| L | 1486 | hCoV-19/Belgium/un-EMC-2/2020\|EPI_ISL_577744\|2020- | ···················· | ···················· | ···················· | ···················· | ···················· | ···················· |
| L | 1487 | hCoV-19/Belgium/un-EMC-4/2020\|EPI_ISL_577746\|2020- | ···················· | ···················· | ···················· | ···················· | ···················· | ···················· |
| L | 1488 | hCoV-19/Belgium/un-EMC-5/2020\|EPI_ISL_577747\|2020- | ···················· | ···················· | ···················· | ···················· | ···················· | ···················· |
| L | 1489 | hCoV-19/mink/Netherlands/NB-EMC-27-3/2020\|EPI_ISL_ | ···················· | ···················· | ···················· | ···················· | ·····T·············· | ···················· |
| L | 1490 | hCoV-19/mink/Netherlands/NB-EMC-28-2/2020\|EPI_ISL_ | ···················· | ···················· | ···················· | ···················· | ···················· | ···················· |
| L | 1491 | hCoV-19/mink/Netherlands/NB-EMC-28-6/2020\|EPI_ISL_ | ···················· | ···················· | ···················· | ···················· | ···················· | ···················· |
| L | 1492 | hCoV-19/mink/Netherlands/NB-EMC-28-7/2020\|EPI_ISL_ | ···················· | ···················· | ···················· | ···················· | ···················· | ···················· |
| L | 1493 | hCoV-19/mink/Netherlands/NB-EMC-28-9/2020\|EPI_ISL_ | ···················· | ···················· | ···················· | ···················· | ···················· | ···················· |
| L | 1494 | hCoV-19/mink/Netherlands/NB-EMC-29-1/2020\|EPI_ISL_ | ···················· | ···················· | ···················· | ···················· | ···················· | ···················· |
| L | 1495 | hCoV-19/mink/Netherlands/NB-EMC-29-10/2020\|EPI_ISL | ···················· | ···················· | ···················· | ···················· | ···················· | ···················· |
| L | 1496 | hCoV-19/mink/Netherlands/NB-EMC-29-20/2020\|EPI_ISL | ···················· | ···················· | ···················· | ···················· | ···················· | ···················· |
| L | 1497 | hCoV-19/mink/Netherlands/NB-EMC-33-1/2020\|EPI_ISL_ | ···················· | ···················· | ···················· | ···················· | ·····T·············· | ···················· |
| L | 1498 | hCoV-19/mink/Netherlands/NB-EMC-33-5/2020\|EPI_ISL_ | ···················· | ···················· | ···················· | ···················· | ·····T·············· | ···················· |
| L | 1499 | hCoV-19/mink/Netherlands/NB-EMC-34-2/2020\|EPI_ISL_ | ···················· | ···················· | ···················· | ···················· | ···················· | ···················· |
| L | 1500 | hCoV-19/mink/Netherlands/NB-EMC-34-4/2020\|EPI_ISL_ | ···················· | ···················· | ···················· | ···················· | ···················· | ···················· |
| L | 1501 | hCoV-19/mink/Netherlands/NB-EMC-34-5/2020\|EPI_ISL_ | ···················· | ···················· | ···················· | ···················· | ···················· | ···················· |
| L | 1502 | hCoV-19/mink/Netherlands/NB-EMC-35-1/2020\|EPI_ISL_ | ···················· | ···················· | ···················· | ···················· | ·····T·············· | ···················· |
| L | 1503 | hCoV-19/mink/Netherlands/NB-EMC-35-2/2020\|EPI_ISL_ | ···················· | ···················· | ···················· | ···················· | ·····T·············· | ···················· |
| L | 1504 | hCoV-19/mink/Netherlands/NB-EMC-35-3/2020\|EPI_ISL_ | ···················· | ···················· | ···················· | ···················· | ·····T·············· | ···················· |
| L | 1505 | hCoV-19/mink/Netherlands/NB-EMC-35-4/2020\|EPI_ISL_ | ···················· | ···················· | ···················· | ···················· | ·····T·············· | ···················· |
| L | 1506 | hCoV-19/mink/Netherlands/NB-EMC-35-5/2020\|EPI_ISL_ | ···················· | ···················· | ···················· | ···················· | ·····T·············· | ···················· |
| L | 1507 | hCoV-19/mink/Netherlands/NB-EMC-36-5/2020\|EPI_ISL_ | ···················· | ···················· | ···················· | ···················· | ·····T·············· | ···················· |
| L | 1508 | hCoV-19/mink/Netherlands/NB-EMC-39-2/2020\|EPI_ISL_ | ···················· | ···················· | ···················· | ···················· | ···················· | ···················· |
| L | 1509 | hCoV-19/mink/Netherlands/NB-EMC-39-3/2020\|EPI_ISL_ | ···················· | ···················· | ···················· | ···················· | ···················· | ···················· |
| L | 1510 | hCoV-19/mink/Netherlands/NB-EMC-39-4/2020\|EPI_ISL_ | ···················· | ···················· | ···················· | ···················· | ···················· | ···················· |
| L | 1511 | hCoV-19/mink/Netherlands/NB-EMC-39-5/2020\|EPI_ISL_ | ···················· | ···················· | ···················· | ···················· | ···················· | ···················· |
| L | 1512 | hCoV-19/mink/Netherlands/NB-EMC-40-3/2020\|EPI_ISL_ | ···················· | ···················· | ···················· | ···················· | ·····T·············· | ···················· |
| L | 1513 | hCoV-19/mink/Netherlands/NB-EMC-41-4/2020\|EPI_ISL_ | ···················· | ···················· | ···················· | ···················· | ···················· | ···················· |
| L | 1514 | hCoV-19/mink/Netherlands/NB-EMC-43-4/2020\|EPI_ISL_ | ···················· | ···················· | ···················· | ···················· | ·····T·············· | ···················· |
| L | 1515 | hCoV-19/mink/Netherlands/NB-EMC-43-5/2020\|EPI_ISL_ | ···················· | ···················· | ···················· | ···················· | ·····T·············· | ···················· |
| L | 1516 | hCoV-19/Canada/MB-NML-166/2020\|EPI_ISL_582243\|2020 | ···················· | ···················· | ···················· | ···················· | ···················· | ···················· |
| L | 1517 | hCoV-19/Canada/MB-NML-167/2020\|EPI_ISL_582244\|2020 | ···················· | ···················· | ···················· | ···················· | ···················· | ···················· |
| L | 1518 | hCoV-19/Canada/MB-NML-168/2020\|EPI_ISL_582245\|2020 | ···················· | ···················· | ···················· | ···················· | ···················· | ···················· |
| L | 1519 | hCoV-19/Canada/MB-NML-409/2020\|EPI_ISL_582246\|2020 | ···················· | ···················· | ···················· | ···················· | ···················· | ···················· |
| L | 1520 | hCoV-19/Canada/MB-NML-410/2020\|EPI_ISL_582247\|2020 | ···················· | ···················· | ···················· | ···················· | ···················· | ···················· |
| L | 1521 | hCoV-19/Canada/MB-NML-411/2020\|EPI_ISL_582248\|2020 | ···················· | ···················· | ···················· | ···················· | ···················· | ···················· |
| L | 1522 | hCoV-19/Canada/MB-NML-412/2020\|EPI_ISL_582249\|2020 | ···················· | ·A·················· | ···················· | ···················· | ···················· | ···················· |
| L | 1523 | hCoV-19/Canada/MB-NML-415/2020\|EPI_ISL_582251\|2020 | ···················· | ···················· | ···················· | ···················· | ···················· | ···················· |
| L | 1524 | hCoV-19/Canada/MB-NML-416/2020\|EPI_ISL_582252\|2020 | ···················· | ···················· | ···················· | ···················· | ···················· | ···················· |
| L | 1525 | hCoV-19/Canada/MB-NML-418/2020\|EPI_ISL_582253\|2020 | ···················· | ···················· | ···················· | ···················· | ···················· | ···················· |
| L | 1526 | hCoV-19/Canada/MB-NML-420/2020\|EPI_ISL_582254\|2020 | ···················· | ···················· | ···················· | ···················· | ···················· | ···················· |
| L | 1527 | hCoV-19/Canada/MB-NML-421/2020\|EPI_ISL_582255\|2020 | ···················· | ···················· | ···················· | ···················· | ···················· | ···················· |
| L | 1528 | hCoV-19/Canada/MB-NML-422/2020\|EPI_ISL_582256\|2020 | ···················· | ···················· | ···················· | ···················· | ···················· | ···················· |
| L | 1529 | hCoV-19/Canada/MB-NML-423/2020\|EPI_ISL_582257\|2020 | ···················· | ···················· | ···················· | ···················· | ···················· | ···················· |
| L | 1530 | hCoV-19/Canada/MB-NML-425/2020\|EPI_ISL_582258\|2020 | ···················· | ···················· | ···················· | ···················· | ···················· | ···················· |
| L | 1531 | hCoV-19/Canada/MB-NML-426/2020\|EPI_ISL_582259\|2020 | ···················· | ···················· | ···················· | ···················· | ···················· | ···················· |
| L | 1532 | hCoV-19/Canada/MB-NML-559/2020\|EPI_ISL_582260\|2020 | ···················· | ···················· | ···················· | ···················· | ···················· | ···················· |
| L | 1533 | hCoV-19/Canada/MB-NML-560/2020\|EPI_ISL_582261\|2020 | ···················· | ···················· | ···················· | ···················· | ···················· | ···················· |
| L | 1534 | hCoV-19/Canada/MB-NML-561/2020\|EPI_ISL_582262\|2020 | ···················· | ···················· | ···················· | ···················· | ···················· | ···················· |
| L | 1535 | hCoV-19/Canada/MB-NML-565/2020\|EPI_ISL_582263\|2020 | ···················· | ···················· | ···················· | ···················· | ···················· | ···················· |
| L | 1536 | hCoV-19/Canada/MB-NML-568/2020\|EPI_ISL_582264\|2020 | ···················· | ···················· | ···················· | ···················· | ···················· | ···················· |
| L | 1537 | hCoV-19/USA/WA-QDX-528/2020\|EPI_ISL_571401\|2020-03 | ···················· | ···················· | ···················· | ···················· | ···················· | ···················· |
| L | 1538 | hCoV-19/USA/WA-QDX-563/2020\|EPI_ISL_571403\|2020-03 | ···················· | ···················· | ···················· | ···················· | ···················· | ···················· |
| L | 1539 | hCoV-19/USA/WA-QDX-564/2020\|EPI_ISL_571404\|2020-03 | ···················· | ···················· | ···················· | ···················· | ···················· | ···················· |
| L | 1540 | hCoV-19/USA/WA-QDX-570/2020\|EPI_ISL_571405\|2020-03 | ···················· | ···················· | ···················· | ···················· | ···················· | ···················· |

| | | | | | | | | |
|---|---|---|---|---|---|---|---|---|
| L | 1541 | hCoV-19/USA/WA-QDX-573/2020\|EPI_ISL_571406\|2020-03 | ···················· | ···················· | ····················· | ···················· | ···················· | ···················· |
| L | 1542 | hCoV-19/USA/WI-QDX-586/2020\|EPI_ISL_571407\|2020-03 | ···················· | ···················· | ····················· | ···················· | ···················· | ···················· |
| L | 1543 | hCoV-19/USA/AR-QDX-679/2020\|EPI_ISL_571408\|2020-03 | ···················· | ···················· | ····················· | ···················· | ···················· | ···················· |
| L | 1544 | hCoV-19/USA/AR-QDX-680/2020\|EPI_ISL_571409\|2020-03 | ···················· | ···················· | ····················· | ···················· | ···················· | ···················· |
| L | 1545 | hCoV-19/USA/CA-QDX-681/2020\|EPI_ISL_571410\|2020-03 | ···················· | ···················· | ····················· | ···················· | ···················· | ···················· |
| L | 1546 | hCoV-19/USA/CA-QDX-692/2020\|EPI_ISL_571411\|2020-03 | ···················· | ···················· | ····················· | ···················· | ···················· | ···················· |
| L | 1547 | hCoV-19/USA/CA-QDX-693/2020\|EPI_ISL_571412\|2020-03 | ···················· | ···················· | ····················· | ···················· | ···················· | ···················· |
| L | 1548 | hCoV-19/USA/CA-QDX-694/2020\|EPI_ISL_571413\|2020-03 | ···················· | ···················· | ····················· | ···················· | ···················· | ···················· |
| L | 1549 | hCoV-19/USA/CA-QDX-695/2020\|EPI_ISL_571414\|2020-03 | ···················· | ···················· | ····················· | ···················· | ···················· | ···················· |
| L | 1550 | hCoV-19/USA/CA-QDX-696/2020\|EPI_ISL_571415\|2020-03 | ···················· | ···················· | ····················· | ···················· | ···················· | ···················· |
| L | 1551 | hCoV-19/USA/CA-QDX-697/2020\|EPI_ISL_571416\|2020-03 | ···················· | ···················· | ····················· | ···················· | ···················· | ···················· |
| L | 1552 | hCoV-19/USA/CA-QDX-698/2020\|EPI_ISL_571417\|2020-03 | ···················· | ···················· | ····················· | ···················· | ···················· | ···················· |
| L | 1553 | hCoV-19/USA/CA-QDX-699/2020\|EPI_ISL_571418\|2020-03 | ···················· | ···················· | ····················· | ···················· | ···················· | ···················· |
| L | 1554 | hCoV-19/USA/CA-QDX-702/2020\|EPI_ISL_571419\|2020-03 | ···················· | ···················· | ····················· | ···················· | ···················· | ···················· |
| L | 1555 | hCoV-19/USA/CA-QDX-703/2020\|EPI_ISL_571420\|2020-03 | ···················· | ···················· | ····················· | ···················· | ···················· | ···················· |
| L | 1556 | hCoV-19/USA/CA-QDX-704/2020\|EPI_ISL_571421\|2020-03 | ···················· | ···················· | ····················· | ···················· | ···················· | ···················· |
| L | 1557 | hCoV-19/USA/CA-QDX-705/2020\|EPI_ISL_571422\|2020-03 | ···················· | ···················· | ····················· | ···················· | ···················· | ···················· |
| L | 1558 | hCoV-19/USA/CA-QDX-706/2020\|EPI_ISL_571423\|2020-03 | ···················· | ···················· | ····················· | ···················· | ···················· | ···················· |
| L | 1559 | hCoV-19/USA/CO-QDX-670/2020\|EPI_ISL_571424\|2020-03 | ···················· | ···················· | ····················· | ···················· | ···················· | ···················· |
| L | 1560 | hCoV-19/USA/CO-QDX-671/2020\|EPI_ISL_571425\|2020-03 | ···················· | ···················· | ····················· | ···················· | ···················· | ···················· |
| L | 1561 | hCoV-19/USA/CO-QDX-672/2020\|EPI_ISL_571426\|2020-03 | ···················· | ···················· | ····················· | ···················· | ···················· | ···················· |
| L | 1562 | hCoV-19/USA/CO-QDX-673/2020\|EPI_ISL_571427\|2020-03 | ···················· | ···················· | ····················· | ···················· | ···················· | ···················· |
| L | 1563 | hCoV-19/USA/CO-QDX-682/2020\|EPI_ISL_571428\|2020-03 | ···················· | ···················· | ····················· | ···················· | ···················· | ···················· |
| L | 1564 | hCoV-19/USA/GA-QDX-684/2020\|EPI_ISL_571429\|2020-03 | ···················· | ···················· | ····················· | ···················· | ···················· | ···················· |
| L | 1565 | hCoV-19/USA/ID-QDX-683/2020\|EPI_ISL_571430\|2020-03 | ···················· | ···················· | ····················· | ···················· | ···················· | ···················· |
| L | 1566 | hCoV-19/USA/ID-QDX-688/2020\|EPI_ISL_571431\|2020-03 | ···················· | ···················· | ····················· | ···················· | ···················· | ···················· |
| L | 1567 | hCoV-19/USA/ID-QDX-700/2020\|EPI_ISL_571432\|2020-03 | ···················· | ···················· | ····················· | ···················· | ···················· | ···················· |
| L | 1568 | hCoV-19/USA/ID-QDX-707/2020\|EPI_ISL_571433\|2020-03 | ···················· | ···················· | ····················· | ···················· | ···················· | ···················· |
| L | 1569 | hCoV-19/USA/IL-QDX-685/2020\|EPI_ISL_571434\|2020-03 | ···················· | ···················· | ····················· | ···················· | ···················· | ···················· |
| L | 1570 | hCoV-19/USA/IL-QDX-686/2020\|EPI_ISL_571435\|2020-03 | ···················· | ···················· | ····················· | ···················· | ···················· | ···················· |
| L | 1571 | hCoV-19/USA/IL-QDX-687/2020\|EPI_ISL_571436\|2020-03 | ···················· | ···················· | ····················· | ···················· | ···················· | ···················· |
| L | 1572 | hCoV-19/USA/ME-QDX-689/2020\|EPI_ISL_571437\|2020-03 | ···················· | ···················· | ····················· | ···················· | ···················· | ···················· |
| L | 1573 | hCoV-19/USA/MI-QDX-690/2020\|EPI_ISL_571438\|2020-03 | ···················· | ···················· | ····················· | ···················· | ···················· | ···················· |
| L | 1574 | hCoV-19/USA/MI-QDX-691/2020\|EPI_ISL_571439\|2020-03 | ···················· | ···················· | ····················· | ···················· | ···················· | ···················· |
| L | 1575 | hCoV-19/USA/MI-QDX-701/2020\|EPI_ISL_571440\|2020-03 | ···················· | ···················· | ····················· | ···················· | ···················· | ···················· |
| L | 1576 | hCoV-19/USA/MI-QDX-708/2020\|EPI_ISL_571441\|2020-03 | ···················· | ···················· | ····················· | ···················· | ···················· | ···················· |
| L | 1577 | hCoV-19/USA/MI-QDX-709/2020\|EPI_ISL_571442\|2020-03 | ···················· | ···················· | ····················· | ···················· | ···················· | ···················· |
| L | 1578 | hCoV-19/USA/MI-QDX-710/2020\|EPI_ISL_571443\|2020-03 | ···················· | ···················· | ····················· | ···················· | ···················· | ···················· |
| L | 1579 | hCoV-19/USA/MI-QDX-711/2020\|EPI_ISL_571444\|2020-03 | ···················· | ···················· | ····················· | ···················· | ···················· | ···················· |
| L | 1580 | hCoV-19/USA/MI-QDX-712/2020\|EPI_ISL_571445\|2020-03 | ···················· | ···················· | ····················· | ···················· | ···················· | ···················· |
| L | 1581 | hCoV-19/USA/MI-QDX-713/2020\|EPI_ISL_571446\|2020-03 | ···················· | ···················· | ····················· | ···················· | ···················· | ···················· |
| L | 1582 | hCoV-19/USA/MI-QDX-714/2020\|EPI_ISL_571447\|2020-03 | ···················· | ···················· | ····················· | ···················· | ···················· | ···················· |
| L | 1583 | hCoV-19/USA/MI-QDX-715/2020\|EPI_ISL_571448\|2020-03 | ···················· | ···················· | ····················· | ···················· | ···················· | ···················· |
| L | 1584 | hCoV-19/USA/MI-QDX-716/2020\|EPI_ISL_571449\|2020-03 | ···················· | ···················· | ····················· | ···················· | ···················· | ···················· |
| L | 1585 | hCoV-19/USA/MI-QDX-717/2020\|EPI_ISL_571450\|2020-03 | ···················· | ···················· | ····················· | ···················· | ···················· | ···················· |
| L | 1586 | hCoV-19/USA/MI-QDX-718/2020\|EPI_ISL_571451\|2020-03 | ···················· | ···················· | ····················· | ···················· | ···················· | ···················· |
| L | 1587 | hCoV-19/USA/CA-QDX-923/2020\|EPI_ISL_571579\|2020-03 | ···················· | ···················· | ····················· | ···················· | ···················· | ···················· |
| L | 1588 | hCoV-19/USA/FL-QDX-911/2020\|EPI_ISL_571595\|2020-03 | ···················· | ···················· | ····················· | ···················· | ···················· | ···················· |
| L | 1589 | hCoV-19/USA/FL-QDX-912/2020\|EPI_ISL_571596\|2020-03 | ···················· | ···················· | ····················· | ···················· | ···················· | ···················· |
| L | 1590 | hCoV-19/USA/FL-QDX-913/2020\|EPI_ISL_571597\|2020-03 | ···················· | ···················· | ····················· | ···················· | ···················· | ···················· |
| L | 1591 | hCoV-19/USA/FL-QDX-943/2020\|EPI_ISL_571598\|2020-03 | ···················· | ···················· | ····················· | ···················· | ···················· | ···················· |
| L | 1592 | hCoV-19/USA/ID-QDX-937/2020\|EPI_ISL_571599\|2020-03 | ···················· | ···················· | ····················· | ···················· | ···················· | ···················· |
| L | 1593 | hCoV-19/England/LIVE-1D1CC7/2020\|EPI_ISL_572710\|20 | ···················· | ···················· | ····················· | ···················· | ···················· | ···················· |
| L | 1595 | hCoV-19/England/MILK-9C72D9/2020\|EPI_ISL_572712\|20 | ···················· | ···················· | ····················· | ···················· | ···················· | ···················· |
| L | 1596 | hCoV-19/England/NORT-2A4CAE/2020\|EPI_ISL_572713\|20 | ···················· | ···················· | ····················· | ···················· | ···················· | ···················· |
| L | 1597 | hCoV-19/England/CAMC-9C1C77/2020\|EPI_ISL_572714\|20 | ···················· | ···················· | ····················· | ···················· | ···················· | ···················· |
| L | 1598 | hCoV-19/England/MILK-9C47CE/2020\|EPI_ISL_572715\|20 | ···················· | ···················· | ····················· | ···················· | ···················· | ···················· |
| L | 1599 | hCoV-19/England/MILK-9C4A4D/2020\|EPI_ISL_572716\|20 | ···················· | ···················· | ····················· | ···················· | ···················· | ···················· |
| L | 1602 | hCoV-19/England/NORW-EF7E4/2020\|EPI_ISL_572719\|202 | ···················· | ···················· | ····················· | ···················· | ···················· | ···················· |
| L | 1604 | hCoV-19/England/MILK-9C495F/2020\|EPI_ISL_572721\|20 | ···················· | ·A·················· | ····················· | ···················· | ···················· | ··A················· |
| L | 1605 | hCoV-19/England/QEUH-9B3A6F/2020\|EPI_ISL_572722\|20 | ···················· | ·A·················· | ····················· | ···················· | ···················· | ··A················· |
| L | 1606 | hCoV-19/England/QEUH-9B9FE2/2020\|EPI_ISL_572723\|20 | ···················· | ·A·················· | ····················· | ···················· | ···················· | ··A················· |
| L | 1607 | hCoV-19/England/QEUH-9BA3A9/2020\|EPI_ISL_572724\|20 | ···················· | ·A·················· | ····················· | ···················· | ···················· | ··A················· |
| L | 1608 | hCoV-19/England/QEUH-9D1B3B/2020\|EPI_ISL_572725\|20 | ···················· | ·A·················· | ····················· | ···················· | ···················· | ··A················· |
| L | 1609 | hCoV-19/England/QEUH-9BA0DF/2020\|EPI_ISL_572726\|20 | ···················· | ···················· | ····················· | ···················· | ···················· | ···················· |
| L | 1610 | hCoV-19/England/QEUH-9B9F88/2020\|EPI_ISL_572727\|20 | ···················· | ·A·················· | ····················· | ···················· | ···················· | ··A················· |
| L | 1611 | hCoV-19/England/QEUH-9BB01A/2020\|EPI_ISL_572728\|20 | ···················· | ·A·················· | ····················· | ···················· | ···················· | ··A················· |
| L | 1612 | hCoV-19/England/NORT-2861B2/2020\|EPI_ISL_572729\|20 | ···················· | ·A·················· | ····················· | ···················· | ···················· | ···················· |
| L | 1613 | hCoV-19/USA/DE-DHSS-F917921/2020\|EPI_ISL_576226\|20 | ···················· | ···················· | ····················· | ···················· | ···················· | ···················· |
| L | 1614 | hCoV-19/USA/DE-DHSS-F917914/2020\|EPI_ISL_576227\|20 | ···················· | ···················· | ····················· | ···················· | ···················· | ···················· |
| L | 1615 | hCoV-19/USA/MN-MDH-1719/2020\|EPI_ISL_576228\|2020-0 | ···················· | ···················· | ····················· | ···················· | ···················· | ···················· |
| L | 1616 | hCoV-19/USA/MN-MDH-1720/2020\|EPI_ISL_576229\|2020-0 | ···················· | ···················· | ····················· | ···················· | ···················· | ···················· |
| L | 1617 | hCoV-19/USA/MN-MDH-1721/2020\|EPI_ISL_576230\|2020-0 | ···················· | ···················· | ····················· | ···················· | ···················· | ···················· |
| L | 1618 | hCoV-19/USA/MN-MDH-1722/2020\|EPI_ISL_576231\|2020-0 | ···················· | ···················· | ····················· | ···················· | ···················· | ···················· |
| L | 1619 | hCoV-19/USA/MN-MDH-1723/2020\|EPI_ISL_576232\|2020-0 | ···················· | ···················· | ····················· | ···················· | ···················· | ···················· |
| L | 1620 | hCoV-19/USA/MN-MDH-1724/2020\|EPI_ISL_576233\|2020-0 | ···················· | ···················· | ····················· | ···················· | ···················· | ···················· |
| L | 1621 | hCoV-19/USA/MN-MDH-1725/2020\|EPI_ISL_576234\|2020-0 | ···················· | ···················· | ····················· | ···················· | ···················· | ···················· |
| L | 1622 | hCoV-19/USA/MN-MDH-1726/2020\|EPI_ISL_576235\|2020-0 | ···················· | ···················· | ····················· | ···················· | ···················· | ···················· |
| L | 1623 | hCoV-19/USA/MN-MDH-1727/2020\|EPI_ISL_576236\|2020-0 | ···················· | ···················· | ····················· | ···················· | ···················· | ···················· |
| L | 1624 | hCoV-19/USA/MN-MDH-1728/2020\|EPI_ISL_576237\|2020-0 | ···················· | ···················· | ····················· | ···················· | ···················· | ···················· |

| | | | | | | | |
|---|---|---|---|---|---|---|---|
| L | 1625 hCoV-19/USA/MN-MDH-1729/2020\|EPI_ISL_576238\|2020-0 | ···················· | ···················· | ···················· | ···················· | ···················· | ···················· |
| L | 1626 hCoV-19/USA/MN-MDH-1730/2020\|EPI_ISL_576239\|2020-0 | ···················· | ···················· | ···················· | ···················· | ···················· | ···················· |
| L | 1627 hCoV-19/USA/MN-MDH-1731/2020\|EPI_ISL_576240\|2020-0 | ···················· | ···················· | ···················· | ···················· | ···················· | ···················· |
| L | 1628 hCoV-19/USA/MN-MDH-1732/2020\|EPI_ISL_576241\|2020-0 | ···················· | ···················· | ···················· | ···················· | ···················· | ···················· |
| L | 1629 hCoV-19/USA/MN-MDH-1733/2020\|EPI_ISL_576242\|2020-0 | ···················· | ···················· | ···················· | ···················· | ···················· | ···················· |
| L | 1630 hCoV-19/USA/MN-MDH-1734/2020\|EPI_ISL_576243\|2020-0 | ···················· | ····G··············· | ···················· | ···················· | ···················· | ···················· |
| L | 1631 hCoV-19/USA/MN-MDH-1735/2020\|EPI_ISL_576244\|2020-0 | ···················· | ···················· | ···················· | ···················· | ···················· | ···················· |
| L | 1632 hCoV-19/USA/MN-MDH-1736/2020\|EPI_ISL_576245\|2020-0 | ···················· | ···················· | ···················· | ···················· | ···················· | ···················· |
| L | 1633 hCoV-19/USA/MN-MDH-1737/2020\|EPI_ISL_576246\|2020-0 | ···················· | ···················· | ···················· | ···················· | ···················· | ···················· |
| L | 1634 hCoV-19/USA/MN-MDH-1738/2020\|EPI_ISL_576247\|2020-0 | ···················· | ···················· | ···················· | ···················· | ···················· | ···················· |
| L | 1635 hCoV-19/USA/MN-MDH-1739/2020\|EPI_ISL_576248\|2020-0 | ···················· | ···················· | ···················· | ···················· | ···················· | ···················· |
| L | 1636 hCoV-19/USA/MN-MDH-1740/2020\|EPI_ISL_576249\|2020-0 | ···················· | ···················· | ···················· | ···················· | ···················· | ···················· |
| L | 1637 hCoV-19/USA/MN-MDH-1741/2020\|EPI_ISL_576250\|2020-0 | ···················· | ···················· | ···················· | ···················· | ···················· | ···················· |
| L | 1638 hCoV-19/USA/MN-MDH-1742/2020\|EPI_ISL_576251\|2020-0 | ···················· | ···················· | ···················· | ···················· | ···················· | ···················· |
| L | 1639 hCoV-19/USA/MN-MDH-1743/2020\|EPI_ISL_576252\|2020-0 | ···················· | ···················· | ···················· | ···················· | ···················· | ···················· |
| L | 1640 hCoV-19/USA/MN-MDH-1744/2020\|EPI_ISL_576253\|2020-0 | ···················· | ···················· | ···················· | ···················· | ···················· | ···················· |
| L | 1641 hCoV-19/USA/MN-MDH-1745/2020\|EPI_ISL_576254\|2020-0 | ···················· | ···················· | ···················· | ···················· | ···················· | ···················· |
| L | 1642 hCoV-19/USA/MN-MDH-1746/2020\|EPI_ISL_576255\|2020-0 | ···················· | ···················· | ···················· | ···················· | ···················· | ···················· |
| L | 1643 hCoV-19/USA/MN-MDH-1747/2020\|EPI_ISL_576256\|2020-0 | ···················· | ···················· | ···················· | ···················· | ···················· | ···················· |
| L | 1644 hCoV-19/Mexico/HID-InDRE-54/2020\|EPI_ISL_576257\|20 | ···················· | ···················· | ···················· | ···················· | ···················· | ···················· |
| L | 1645 hCoV-19/Mexico/AGU-InDRE-55/2020\|EPI_ISL_576258\|20 | ···················· | ·A·················· | ···················· | ···················· | ···················· | ···················· |
| L | 1646 hCoV-19/Mexico/TLA-InDRE-56/2020\|EPI_ISL_576259\|20 | ········T··········· | ···················· | ···················· | ···················· | ···················· | ···················· |
| L | 1647 hCoV-19/Mexico/TLA-InDRE-57/2020\|EPI_ISL_576260\|20 | ···················· | ···················· | ···················· | ···················· | ···················· | ···················· |
| L | 1648 hCoV-19/Mexico/AGU-InDRE-58/2020\|EPI_ISL_576261\|20 | ···················· | ···················· | ···················· | ···················· | ···················· | ···················· |
| L | 1649 hCoV-19/Mexico/NLE-InDRE-59/2020\|EPI_ISL_576262\|20 | ···················· | ···················· | ···················· | ···················· | ···················· | ···················· |
| L | 1650 hCoV-19/Mexico/NLE-InDRE-60/2020\|EPI_ISL_576263\|20 | ···················· | ···················· | ···················· | ···················· | ···················· | ···················· |
| L | 1651 hCoV-19/Mexico/OAX-InDRE-61/2020\|EPI_ISL_576264\|20 | ···················· | ·A·················· | ···················· | ···················· | ···················· | ···················· |
| L | 1652 hCoV-19/Mexico/CAM-InDRE-62/2020\|EPI_ISL_576265\|20 | ···················· | ·A·················· | ···················· | ···················· | ···················· | ···················· |
| L | 1653 hCoV-19/Mexico/CAM-InDRE-63/2020\|EPI_ISL_576266\|20 | ···················· | ···················· | ···················· | ···················· | ···················· | ···················· |
| L | 1654 hCoV-19/Mexico/CAM-InDRE-64/2020\|EPI_ISL_576267\|20 | ···················· | ···················· | ···················· | ···················· | ···················· | ···················· |
| L | 1655 hCoV-19/Mexico/HID-InDRE-65/2020\|EPI_ISL_576268\|20 | ···················· | ···················· | ···················· | ···················· | ···················· | ···················· |
| L | 1656 hCoV-19/Mexico/VER-InDRE-66/2020\|EPI_ISL_576269\|20 | ···················· | ···················· | ···················· | ···················· | ···················· | ···················· |
| L | 1657 hCoV-19/Mexico/VER-InDRE-67/2020\|EPI_ISL_576270\|20 | ···················· | ···················· | ···················· | ···················· | ···················· | ···················· |
| L | 1658 hCoV-19/Mexico/MIC-InDRE-68/2020\|EPI_ISL_576271\|20 | ···················· | ···················· | ···················· | ···················· | ···················· | ···················· |
| L | 1659 hCoV-19/Mexico/SLP-InDRE-69/2020\|EPI_ISL_576272\|20 | ···················· | ·A·················· | ···················· | ···················· | ···················· | ···················· |
| L | 1660 hCoV-19/Mexico/ZAC-InDRE-70/2020\|EPI_ISL_576273\|20 | ···················· | ·A·················· | ···················· | ···················· | ···················· | ···················· |
| L | 1661 hCoV-19/Mexico/ZAC-InDRE-71/2020\|EPI_ISL_576274\|20 | ···················· | ·A·················· | ···················· | ···················· | ···················· | ···················· |
| L | 1662 hCoV-19/Mexico/ZAC-InDRE-72/2020\|EPI_ISL_576275\|20 | ···················· | ···················· | ···················· | ···················· | ···················· | ···················· |
| L | 1663 hCoV-19/Mexico/MIC-InDRE-73/2020\|EPI_ISL_576276\|20 | ···················· | ·A·················· | ···················· | ···················· | ···················· | ···················· |
| L | 1664 hCoV-19/Mexico/MIC-InDRE-74/2020\|EPI_ISL_576277\|20 | ···················· | ···················· | ···················· | ···················· | ···················· | ···················· |
| L | 1665 hCoV-19/Mexico/COL-InDRE-75/2020\|EPI_ISL_576278\|20 | ···················· | ·A·················· | ···················· | ···················· | ···················· | ···················· |
| L | 1666 hCoV-19/Mexico/COL-InDRE-76/2020\|EPI_ISL_576279\|20 | ···················· | ·A·················· | ···················· | ···················· | ···················· | ···················· |
| L | 1667 hCoV-19/USA/MI-MDHHS-SC21999/2020\|EPI_ISL_577565\|2 | ···················· | ···················· | ···················· | ···················· | ···················· | ···················· |
| L | 1668 hCoV-19/USA/MI-MDHHS-SC22001/2020\|EPI_ISL_577566\|2 | ···················· | ···················· | ···················· | ···················· | ···················· | ···················· |
| L | 1669 hCoV-19/USA/MI-MDHHS-SC22002/2020\|EPI_ISL_577567\|2 | ···················· | ·A·················· | ···················· | ···················· | ···················· | ···················· |
| L | 1670 hCoV-19/USA/MI-MDHHS-SC22006/2020\|EPI_ISL_577568\|2 | ···················· | ·············A······ | ···················· | ···················· | ···················· | ···················· |
| L | 1671 hCoV-19/USA/MI-MDHHS-SC22007/2020\|EPI_ISL_577569\|2 | ···················· | ···················· | ···················· | ···················· | ···················· | ···················· |
| L | 1672 hCoV-19/USA/MI-MDHHS-SC22008/2020\|EPI_ISL_577570\|2 | ···················· | ·············A······ | ···················· | ···················· | ···················· | ···················· |
| L | 1673 hCoV-19/USA/MI-MDHHS-SC22009/2020\|EPI_ISL_577571\|2 | ···················· | ···················· | ···················· | ···················· | ···················· | ···················· |
| L | 1674 hCoV-19/USA/MI-MDHHS-SC22010/2020\|EPI_ISL_577572\|2 | ···················· | ···················· | ···················· | ···················· | ···················· | ···················· |
| L | 1675 hCoV-19/USA/MI-MDHHS-SC22012/2020\|EPI_ISL_577573\|2 | ···················· | ·············A······ | ···················· | ···················· | ···················· | ···················· |
| L | 1676 hCoV-19/USA/MI-MDHHS-SC22013/2020\|EPI_ISL_577574\|2 | ···················· | ···················· | ···················· | ···················· | ···················· | ···················· |
| L | 1677 hCoV-19/USA/MI-MDHHS-SC22015/2020\|EPI_ISL_577575\|2 | ···················· | ···················· | ···················· | ···················· | ···················· | ···················· |
| L | 1678 hCoV-19/USA/MI-MDHHS-SC22016/2020\|EPI_ISL_577576\|2 | ···················· | ···················· | ···················· | ···················· | ···················· | ···················· |
| L | 1679 hCoV-19/USA/MI-MDHHS-SC22017/2020\|EPI_ISL_577577\|2 | ···················· | ···················· | ···················· | ···················· | ···················· | ···················· |
| L | 1680 hCoV-19/USA/MI-MDHHS-SC22019/2020\|EPI_ISL_577578\|2 | ···················· | ·A····A············· | ···················· | ···················· | ···················· | ···················· |
| L | 1681 hCoV-19/USA/MI-MDHHS-SC22020/2020\|EPI_ISL_577579\|2 | ···················· | ···················· | ···················· | ···················· | ···················· | ···················· |
| L | 1682 hCoV-19/USA/MI-MDHHS-SC22024/2020\|EPI_ISL_577580\|2 | ···················· | ···················· | ···················· | ···················· | ···················· | ···················· |
| L | 1683 hCoV-19/USA/MI-MDHHS-SC22025/2020\|EPI_ISL_577581\|2 | ···················· | ·A·················· | ···················· | ···················· | ···················· | ···················· |
| L | 1684 hCoV-19/USA/MI-MDHHS-SC22026/2020\|EPI_ISL_577582\|2 | ···················· | ···················· | ···················· | ···················· | ···················· | ···················· |
| L | 1685 hCoV-19/USA/MI-MDHHS-SC22027/2020\|EPI_ISL_577583\|2 | ···················· | ···················· | ···················· | ···················· | ···················· | ···················· |
| L | 1686 hCoV-19/USA/MI-MDHHS-SC22028/2020\|EPI_ISL_577584\|2 | ···················· | ···················· | ···················· | ···················· | ···················· | ···················· |
| L | 1687 hCoV-19/USA/MI-MDHHS-SC22029/2020\|EPI_ISL_577585\|2 | ···················· | ···················· | ···················· | ···················· | ···················· | ···················· |
| L | 1688 hCoV-19/USA/MI-MDHHS-SC22030/2020\|EPI_ISL_577586\|2 | ···················· | ·A·················· | ···················· | ···················· | ···················· | ···················· |
| L | 1689 hCoV-19/USA/MI-MDHHS-SC22031/2020\|EPI_ISL_577587\|2 | ···················· | ···················· | ···················· | ···················· | ···················· | ···················· |
| L | 1690 hCoV-19/USA/MI-MDHHS-SC22036/2020\|EPI_ISL_577588\|2 | ···················· | ···················· | ···················· | ···················· | ···················· | ···················· |
| L | 1691 hCoV-19/USA/MI-MDHHS-SC22038/2020\|EPI_ISL_577589\|2 | ···················· | ···················· | ···················· | ···················· | ···················· | ···················· |
| L | 1692 hCoV-19/USA/MI-MDHHS-SC22039/2020\|EPI_ISL_577590\|2 | ···················· | ···················· | ···················· | ···················· | ···················· | ···················· |
| L | 1693 hCoV-19/USA/MI-MDHHS-SC22041/2020\|EPI_ISL_577591\|2 | ···················· | ···················· | ···················· | ···················· | ···················· | ···················· |
| L | 1694 hCoV-19/USA/MI-MDHHS-SC22042/2020\|EPI_ISL_577592\|2 | ···················· | ···················· | ···················· | ···················· | ···················· | ···················· |
| L | 1695 hCoV-19/USA/MI-MDHHS-SC22043/2020\|EPI_ISL_577593\|2 | ···················· | ···················· | ···················· | ···················· | ···················· | ···················· |
| L | 1696 hCoV-19/USA/MI-MDHHS-SC22045/2020\|EPI_ISL_577594\|2 | ···················· | ···················· | ···················· | ···················· | ···················· | ···················· |
| L | 1697 hCoV-19/USA/MI-MDHHS-SC22046/2020\|EPI_ISL_577595\|2 | ···················· | ·A·················· | ···················· | ···················· | ···················· | ···················· |
| L | 1698 hCoV-19/USA/MI-MDHHS-SC22047/2020\|EPI_ISL_577596\|2 | ···················· | ···················· | ···················· | ···················· | ···················· | ···················· |
| L | 1699 hCoV-19/Australia/NSW2827/2020\|EPI_ISL_577597\|2020 | ···················· | ····A··············· | ···················· | ···················· | ···················· | ···················· |
| L | 1700 hCoV-19/Australia/TAS00279/2020\|EPI_ISL_577607\|202 | ···················· | ···················· | ···················· | ···················· | ···················· | ···················· |
| L | 1701 hCoV-19/India/MH-NIV-229720/2020\|EPI_ISL_577700\|20 | ···················· | ···················· | ···················· | ···················· | ···················· | ···················· |
| L | 1702 hCoV-19/India/MH-NIV-229771/2020\|EPI_ISL_577701\|20 | ···················· | ···················· | ···················· | ···················· | ···················· | ···················· |
| L | 1703 hCoV-19/India/MH-NIV-229912/2020\|EPI_ISL_577702\|20 | ···················· | ·A·················· | ···················· | ···················· | ···················· | ···A················ |
| L | 1704 hCoV-19/India/MH-NIV-206715/2020\|EPI_ISL_577704\|20 | ···················· | ···················· | ···················· | ···················· | ···················· | ···················· |

| | | | | | | | |
|---|---|---|---|---|---|---|---|
| L | 1705 hCoV-19/India/MH-NIV-211414/2020\|EPI_ISL_577705\|20 | ···················· | ···················· | ····················· | ···················· | ···················· | ···················· |
| L | 1706 hCoV-19/India/MH-NIV-QC-1459/2020\|EPI_ISL_577707\|2 | ···················· | ···················· | ····················· | ···················· | ···················· | ···················· |
| L | 1707 hCoV-19/India/MH-NIV-229898/2020\|EPI_ISL_577709\|20 | ···················· | ···················· | ····················· | ···················· | ···················· | ···················· |
| L | 1708 hCoV-19/India/MH-NIV-229297/2020\|EPI_ISL_577710\|20 | ···················· | ···················· | ····················· | ···················· | ···················· | ···················· |
| L | 1709 hCoV-19/USA/LA-EVTL484/2020\|EPI_ISL_578888\|2020-06 | ···················· | ···················· | ····················· | ···················· | ···················· | ···················· |
| L | 1710 hCoV-19/USA/LA-EVTL485/2020\|EPI_ISL_578889\|2020-07 | ···················· | ······A············· | ····················· | ···················· | ···················· | ···················· |
| L | 1711 hCoV-19/USA/LA-EVTL486/2020\|EPI_ISL_578890\|2020-06 | ···················· | ····A··············· | ····················· | ···················· | ···················· | ···················· |
| L | 1712 hCoV-19/USA/LA-EVTL487/2020\|EPI_ISL_578891\|2020-07 | ···················· | ······A············· | ····················· | ···················· | ···················· | ···················· |
| L | 1713 hCoV-19/USA/LA-EVTL488/2020\|EPI_ISL_578892\|2020-06 | ···················· | ···················· | ····················· | ···················· | ···················· | ···················· |
| L | 1714 hCoV-19/USA/LA-EVTL489/2020\|EPI_ISL_578893\|2020-07 | ···················· | ···················· | ····················· | ···················· | ···················· | ···················· |
| L | 1715 hCoV-19/USA/LA-EVTL490/2020\|EPI_ISL_578894\|2020-06 | ···················· | ···················· | ····················· | ···················· | ···················· | ···················· |
| L | 1716 hCoV-19/USA/LA-EVTL491/2020\|EPI_ISL_578895\|2020-07 | ···················· | ······A············· | ····················· | ···················· | ···················· | ···················· |
| L | 1717 hCoV-19/USA/LA-EVTL492/2020\|EPI_ISL_578896\|2020-07 | ···················· | ······A············· | ····················· | ···················· | ···················· | ···················· |
| L | 1718 hCoV-19/USA/LA-EVTL493/2020\|EPI_ISL_578897\|2020-07 | ···················· | ······A············· | ····················· | ···················· | ···················· | ···················· |
| L | 1719 hCoV-19/USA/LA-EVTL494/2020\|EPI_ISL_578898\|2020-07 | ···················· | ···················· | ····················· | ···················· | ···················· | ···················· |
| L | 1720 hCoV-19/USA/LA-EVTL495/2020\|EPI_ISL_578899\|2020-06 | ···················· | ···················· | ····················· | ···················· | ···················· | ···················· |
| L | 1721 hCoV-19/USA/LA-EVTL528/2020\|EPI_ISL_578932\|2020-06 | ···················· | ···················· | ····················· | ···················· | ·····C·············· | ···················· |
| L | 1722 hCoV-19/USA/LA-EVTL529/2020\|EPI_ISL_578933\|2020-08 | ···················· | ·A·················· | ····················· | ···················· | ···················· | ···················· |
| L | 1723 hCoV-19/USA/LA-EVTL532/2020\|EPI_ISL_578936\|2020-06 | ···················· | ···················· | ····················· | ···················· | ···················· | ···················· |
| L | 1724 hCoV-19/USA/LA-EVTL533/2020\|EPI_ISL_578937\|2020-08 | ···················· | ···················· | ····················· | ···················· | ···················· | ···················· |
| L | 1725 hCoV-19/USA/LA-EVTL534/2020\|EPI_ISL_578938\|2020-08 | ···················· | ···················· | ····················· | ···················· | ···················· | ···················· |
| L | 1726 hCoV-19/USA/LA-EVTL535/2020\|EPI_ISL_578939\|2020-08 | ···················· | ···················· | ····················· | ···················· | ···················· | ···················· |
| L | 1727 hCoV-19/USA/LA-EVTL536/2020\|EPI_ISL_578940\|2020-06 | ···················· | ···················· | ····················· | ···················· | ···················· | ···················· |
| L | 1728 hCoV-19/USA/LA-EVTL537/2020\|EPI_ISL_578941\|2020-06 | ···················· | ···················· | ····················· | ···················· | ···················· | ···················· |
| L | 1729 hCoV-19/USA/LA-EVTL538/2020\|EPI_ISL_578942\|2020-06 | ···················· | ···················· | ····················· | ···················· | ···················· | ···················· |
| L | 1730 hCoV-19/India/MH-NIV-229543/2020\|EPI_ISL_582241\|20 | ···················· | ···················· | ····················· | ···················· | ···················· | ···················· |
| L | 1731 hCoV-19/Canada/MB-NML-163/2020\|EPI_ISL_582242\|2020 | ···················· | ···················· | ····················· | ···················· | ···················· | ···················· |
| L | 1732 hCoV-19/Canada/MB-NML-569/2020\|EPI_ISL_582265\|2020 | ···················· | ···················· | ····················· | ···················· | ···················· | ···················· |
| L | 1733 hCoV-19/Canada/MB-NML-571/2020\|EPI_ISL_582266\|2020 | ···················· | ···················· | ····················· | ···················· | ···················· | ···················· |
| L | 1734 hCoV-19/Canada/MB-NML-572/2020\|EPI_ISL_582267\|2020 | ···················· | ···················· | ····················· | ···················· | ···················· | ···················· |
| L | 1735 hCoV-19/Canada/MB-NML-573/2020\|EPI_ISL_582268\|2020 | ···················· | ···················· | ····················· | ···················· | ···················· | ···················· |
| L | 1736 hCoV-19/Canada/MB-NML-574/2020\|EPI_ISL_582269\|2020 | ···················· | ···················· | ····················· | ···················· | ···················· | ···················· |
| L | 1737 hCoV-19/Canada/MB-NML-576/2020\|EPI_ISL_582270\|2020 | ···················· | ···················· | ····················· | ···················· | ···················· | ···················· |
| L | 1738 hCoV-19/Canada/MB-NML-580/2020\|EPI_ISL_582271\|2020 | ···················· | ···················· | ····················· | ···················· | ···················· | ···················· |
| L | 1739 hCoV-19/Canada/MB-NML-581/2020\|EPI_ISL_582272\|2020 | ···················· | ···················· | ····················· | ···················· | ···················· | ···················· |
| L | 1740 hCoV-19/Canada/MB-NML-583/2020\|EPI_ISL_582273\|2020 | ···················· | ···················· | ····················· | ···················· | ···················· | ···················· |
| L | 1741 hCoV-19/Canada/MB-NML-584/2020\|EPI_ISL_582274\|2020 | ···················· | ···················· | ····················· | ···················· | ···················· | ···················· |
| L | 1742 hCoV-19/Canada/MB-NML-585/2020\|EPI_ISL_582275\|2020 | ···················· | ···················· | ····················· | ···················· | ···················· | ···················· |
| L | 1743 hCoV-19/Canada/MB-NML-587/2020\|EPI_ISL_582276\|2020 | ···················· | ···················· | ····················· | ···················· | ···················· | ···················· |
| L | 1744 hCoV-19/Canada/MB-NML-590/2020\|EPI_ISL_582277\|2020 | ···················· | ···················· | ····················· | ···················· | ···················· | ···················· |
| L | 1745 hCoV-19/Canada/MB-NML-591/2020\|EPI_ISL_582278\|2020 | ···················· | ·A·················· | ····················· | ···················· | ···················· | ···················· |
| L | 1746 hCoV-19/Canada/MB-NML-592/2020\|EPI_ISL_582279\|2020 | ···················· | ···················· | ····················· | ···················· | ···················· | ···················· |
| L | 1747 hCoV-19/Canada/MB-NML-593/2020\|EPI_ISL_582280\|2020 | ···················· | ···················· | ····················· | ···················· | ···················· | ···················· |
| L | 1748 hCoV-19/Canada/MB-NML-594/2020\|EPI_ISL_582281\|2020 | ···················· | ···················· | ····················· | ···················· | ···················· | ···················· |
| L | 1749 hCoV-19/Canada/MB-NML-597/2020\|EPI_ISL_582282\|2020 | ···················· | ···················· | ····················· | ···················· | ···················· | ···················· |
| L | 1750 hCoV-19/Canada/MB-NML-598/2020\|EPI_ISL_582283\|2020 | ···················· | ···················· | ····················· | ···················· | ···················· | ···················· |
| L | 1751 hCoV-19/Canada/MB-NML-600/2020\|EPI_ISL_582284\|2020 | ···················· | ···················· | ····················· | ···················· | ···················· | ···················· |
| L | 1752 hCoV-19/Canada/MB-NML-603/2020\|EPI_ISL_582285\|2020 | ···················· | ···················· | ····················· | ···················· | ···················· | ···················· |
| L | 1753 hCoV-19/Canada/MB-NML-612/2020\|EPI_ISL_582286\|2020 | ···················· | ···················· | ····················· | ···················· | ···················· | ···················· |
| L | 1754 hCoV-19/Canada/MB-NML-616/2020\|EPI_ISL_582288\|2020 | ···················· | ···················· | ····················· | ···················· | ···················· | ···················· |
| L | 1755 hCoV-19/Canada/MB-NML-617/2020\|EPI_ISL_582289\|2020 | ···················· | ···················· | ····················· | ···················· | ···················· | ···················· |
| L | 1756 hCoV-19/Canada/MB-NML-619/2020\|EPI_ISL_582290\|2020 | ···················· | ···················· | ····················· | ···················· | ···················· | ···················· |
| L | 1757 hCoV-19/Canada/MB-NML-621/2020\|EPI_ISL_582291\|2020 | ···················· | ···················· | ····················· | ···················· | ···················· | ···················· |
| L | 1758 hCoV-19/Canada/MB-NML-623/2020\|EPI_ISL_582292\|2020 | ···················· | ···················· | ····················· | ···················· | ···················· | ···················· |
| L | 1759 hCoV-19/Canada/MB-NML-626/2020\|EPI_ISL_582293\|2020 | ···················· | ···················· | ····················· | ···················· | ···················· | ···················· |
| L | 1760 hCoV-19/Canada/MB-NML-628/2020\|EPI_ISL_582294\|2020 | ···················· | ···················· | ····················· | ···················· | ···················· | ···················· |
| L | 1761 hCoV-19/Canada/MB-NML-629/2020\|EPI_ISL_582295\|2020 | ···················· | ···················· | ····················· | ···················· | ···················· | ···················· |
| L | 1762 hCoV-19/Canada/MB-NML-636/2020\|EPI_ISL_582296\|2020 | ···················· | ···················· | ····················· | ···················· | ···················· | ···················· |
| L | 1763 hCoV-19/Canada/MB-NML-640/2020\|EPI_ISL_582297\|2020 | ···················· | ···················· | ····················· | ···················· | ···················· | ···················· |
| L | 1764 hCoV-19/Canada/MB-NML-641/2020\|EPI_ISL_582298\|2020 | ···················· | ···················· | ····················· | ···················· | ···················· | ···················· |
| L | 1765 hCoV-19/Canada/MB-NML-642/2020\|EPI_ISL_582299\|2020 | ···················· | ···················· | ····················· | ···················· | ···················· | ···················· |
| L | 1767 hCoV-19/USA/MI-UM-10035884804/2020\|EPI_ISL_583401\| | ···················· | ···················· | ····················· | ···················· | ···················· | ···················· |
| L | 1772 hCoV-19/USA/MI-UM-10036064450/2020\|EPI_ISL_583407\| | ···················· | ·A·················· | ····················· | ···················· | ···················· | ···················· |
| L | 1773 hCoV-19/USA/MI-UM-10036079833/2020\|EPI_ISL_583408\| | ···················· | ···················· | ····················· | ···················· | ···················· | ···················· |
| L | 1777 hCoV-19/USA/MI-UM-10036061974/2020\|EPI_ISL_583413\| | ···················· | ···················· | ····················· | ···················· | ···················· | ···················· |
| L | 1783 hCoV-19/USA/MI-UM-10036074663/2020\|EPI_ISL_583419\| | ···················· | ·A·················· | ····················· | ···················· | ···················· | ···················· |
| L | 1787 hCoV-19/USA/MI-UM-10036100373/2020\|EPI_ISL_583424\| | ···················· | ···················· | ····················· | ···················· | ···················· | ···················· |
| L | 1789 hCoV-19/USA/NY-MSK-1Ai/2020\|EPI_ISL_583426\|2020-04 | ···················· | ···················· | ····················· | ···················· | ···················· | ···················· |
| L | 1790 hCoV-19/USA/NY-MSK-1A/2020\|EPI_ISL_583427\|2020-04- | ···················· | ···················· | ····················· | ···················· | ···················· | ···················· |
| L | 1791 hCoV-19/USA/NY-MSK-2A/2020\|EPI_ISL_583428\|2020-03- | ···················· | ···················· | ····················· | ···················· | ···················· | ···················· |
| L | 1792 hCoV-19/USA/NY-MSK-2Ai/2020\|EPI_ISL_583429\|2020-03 | ···················· | ···················· | ····················· | ···················· | ···················· | ···················· |
| L | 1793 hCoV-19/USA/MI-QDX-719/2020\|EPI_ISL_571452\|2020-03 | ···················· | ···················· | ····················· | ···················· | ···················· | ···················· |
| L | 1794 hCoV-19/USA/MI-QDX-720/2020\|EPI_ISL_571453\|2020-03 | ···················· | ···················· | ····················· | ···················· | ···················· | ···················· |
| L | 1795 hCoV-19/USA/MI-QDX-721/2020\|EPI_ISL_571454\|2020-03 | ···················· | ···················· | ····················· | ···················· | ···················· | ···················· |
| L | 1796 hCoV-19/USA/MI-QDX-722/2020\|EPI_ISL_571455\|2020-03 | ···················· | ···················· | ····················· | ···················· | ···················· | ···················· |
| L | 1797 hCoV-19/USA/MI-QDX-723/2020\|EPI_ISL_571456\|2020-03 | ···················· | ···················· | ····················· | ···················· | ···················· | ···················· |
| L | 1798 hCoV-19/USA/MI-QDX-724/2020\|EPI_ISL_571457\|2020-03 | ···················· | ···················· | ····················· | ···················· | ···················· | ···················· |
| L | 1799 hCoV-19/USA/MI-QDX-725/2020\|EPI_ISL_571458\|2020-03 | ···················· | ···················· | ····················· | ···················· | ···················· | ···················· |
| L | 1800 hCoV-19/USA/MI-QDX-726/2020\|EPI_ISL_571459\|2020-03 | ···················· | ···················· | ····················· | ···················· | ···················· | ···················· |
| L | 1801 hCoV-19/USA/MI-QDX-727/2020\|EPI_ISL_571460\|2020-03 | ···················· | ···················· | ····················· | ···················· | ···················· | ···················· |

| | | | | | | | | |
|---|---|---|---|---|---|---|---|---|
| L | 1802 hCoV-19/USA/MI-QDX-728/2020\|EPI_ISL_571461\|2020-03 | ···················· | ···················· | ····················· | ···················· | ···················· | ···················· | |
| L | 1803 hCoV-19/USA/MI-QDX-729/2020\|EPI_ISL_571462\|2020-03 | ···················· | ···················· | ····················· | ···················· | ···················· | ···················· | |
| L | 1804 hCoV-19/USA/MI-QDX-730/2020\|EPI_ISL_571463\|2020-03 | ···················· | ···················· | ····················· | ···················· | ···················· | ···················· | |
| L | 1805 hCoV-19/USA/MI-QDX-731/2020\|EPI_ISL_571464\|2020-03 | ···················· | ···················· | ····················· | ···················· | ···················· | ···················· | |
| L | 1806 hCoV-19/USA/MI-QDX-732/2020\|EPI_ISL_571465\|2020-03 | ···················· | ···················· | ····················· | ···················· | ···················· | ···················· | |
| L | 1807 hCoV-19/USA/MI-QDX-733/2020\|EPI_ISL_571466\|2020-03 | ···················· | ···················· | ····················· | ···················· | ···················· | ···················· | |
| L | 1808 hCoV-19/USA/MI-QDX-734/2020\|EPI_ISL_571467\|2020-03 | ···················· | ···················· | ····················· | ···················· | ···················· | ···················· | |
| L | 1809 hCoV-19/USA/MI-QDX-735/2020\|EPI_ISL_571468\|2020-03 | ···················· | ···················· | ····················· | ···················· | ···················· | ···················· | |
| L | 1810 hCoV-19/USA/MI-QDX-736/2020\|EPI_ISL_571469\|2020-03 | ···················· | ···················· | ····················· | ···················· | ···················· | ···················· | |
| L | 1811 hCoV-19/USA/MI-QDX-737/2020\|EPI_ISL_571470\|2020-03 | ···················· | ···················· | ····················· | ···················· | ···················· | ···················· | |
| L | 1812 hCoV-19/USA/MI-QDX-738/2020\|EPI_ISL_571471\|2020-03 | ···················· | ···················· | ····················· | ···················· | ···················· | ···················· | |
| L | 1813 hCoV-19/USA/MI-QDX-739/2020\|EPI_ISL_571472\|2020-03 | ···················· | ···················· | ····················· | ···················· | ···················· | ···················· | |
| L | 1814 hCoV-19/USA/MO-QDX-674/2020\|EPI_ISL_571473\|2020-03 | ···················· | ···················· | ····················· | ···················· | ···················· | ···················· | |
| L | 1815 hCoV-19/USA/MO-QDX-675/2020\|EPI_ISL_571474\|2020-03 | ···················· | ···················· | ····················· | ···················· | ···················· | ···················· | |
| L | 1816 hCoV-19/USA/MO-QDX-676/2020\|EPI_ISL_571475\|2020-03 | ···················· | ···················· | ····················· | ···················· | ···················· | ···················· | |
| L | 1817 hCoV-19/USA/MO-QDX-677/2020\|EPI_ISL_571476\|2020-03 | ···················· | ···················· | ····················· | ···················· | ···················· | ···················· | |
| L | 1818 hCoV-19/USA/MO-QDX-678/2020\|EPI_ISL_571477\|2020-03 | ···················· | ···················· | ····················· | ···················· | ···················· | ···················· | |
| L | 1819 hCoV-19/USA/TX-QDX-664/2020\|EPI_ISL_571478\|2020-03 | ···················· | ···················· | ····················· | ···················· | ···················· | ···················· | |
| L | 1820 hCoV-19/USA/TX-QDX-665/2020\|EPI_ISL_571479\|2020-03 | ···················· | ···················· | ····················· | ···················· | ···················· | ···················· | |
| L | 1821 hCoV-19/USA/TX-QDX-666/2020\|EPI_ISL_571480\|2020-03 | ···················· | ···················· | ····················· | ···················· | ···················· | ···················· | |
| L | 1822 hCoV-19/USA/TX-QDX-667/2020\|EPI_ISL_571481\|2020-03 | ···················· | ···················· | ····················· | ···················· | ···················· | ···················· | |
| L | 1823 hCoV-19/USA/TX-QDX-668/2020\|EPI_ISL_571482\|2020-03 | ···················· | ···················· | ····················· | ···················· | ···A················ | ···················· | |
| L | 1824 hCoV-19/USA/TX-QDX-669/2020\|EPI_ISL_571483\|2020-03 | ···················· | ···················· | ····················· | ···················· | ···················· | ···················· | |
| L | 1825 hCoV-19/USA/CA-QDX-774/2020\|EPI_ISL_571484\|2020-03 | ···················· | ···················· | ····················· | ···················· | ···················· | ···················· | |
| L | 1826 hCoV-19/USA/CA-QDX-776/2020\|EPI_ISL_571486\|2020-03 | ···················· | ···················· | ····················· | ···················· | ···················· | ···················· | |
| L | 1827 hCoV-19/USA/CA-QDX-781/2020\|EPI_ISL_571487\|2020-03 | ···················· | ···················· | ····················· | ···················· | ···················· | ···················· | |
| L | 1828 hCoV-19/USA/CA-QDX-794/2020\|EPI_ISL_571488\|2020-03 | ···················· | ···················· | ····················· | ···················· | ···················· | ···················· | |
| L | 1829 hCoV-19/USA/CA-QDX-795/2020\|EPI_ISL_571489\|2020-03 | ···················· | ···················· | ····················· | ···················· | ···················· | ···················· | |
| L | 1830 hCoV-19/USA/CA-QDX-796/2020\|EPI_ISL_571490\|2020-03 | ···················· | ···················· | ····················· | ···················· | ···················· | ···················· | |
| L | 1831 hCoV-19/USA/CA-QDX-797/2020\|EPI_ISL_571491\|2020-03 | ···················· | ···················· | ····················· | ···················· | ···················· | ···················· | |
| L | 1832 hCoV-19/USA/CA-QDX-799/2020\|EPI_ISL_571492\|2020-03 | ···················· | ···················· | ····················· | ···················· | ···················· | ···················· | |
| L | 1833 hCoV-19/USA/CA-QDX-800/2020\|EPI_ISL_571493\|2020-03 | ···················· | ···················· | ····················· | ···················· | ···················· | ···················· | |
| L | 1834 hCoV-19/USA/CA-QDX-801/2020\|EPI_ISL_571494\|2020-03 | ···················· | ···················· | ····················· | ···················· | ···················· | ···················· | |
| L | 1835 hCoV-19/USA/CA-QDX-802/2020\|EPI_ISL_571495\|2020-03 | ···················· | ···················· | ····················· | ···················· | ···················· | ···················· | |
| L | 1836 hCoV-19/USA/CA-QDX-803/2020\|EPI_ISL_571496\|2020-03 | ···················· | ···················· | ····················· | ···················· | ···················· | ···················· | |
| L | 1837 hCoV-19/USA/CA-QDX-804/2020\|EPI_ISL_571497\|2020-03 | ···················· | ···················· | ····················· | ···················· | ···················· | ···················· | |
| L | 1838 hCoV-19/USA/CA-QDX-805/2020\|EPI_ISL_571498\|2020-03 | ···················· | ···················· | ····················· | ···················· | ···················· | ···················· | |
| L | 1839 hCoV-19/USA/CA-QDX-806/2020\|EPI_ISL_571499\|2020-03 | ···················· | ···················· | ····················· | ···················· | ···················· | ···················· | |
| L | 1840 hCoV-19/Switzerland/VD-ETHZ-100240/2020\|EPI_ISL_57 | ···················· | ···················· | ····················· | ···················· | ···················· | ···················· | |
| L | 1841 hCoV-19/Switzerland/VD-ETHZ-100302/2020\|EPI_ISL_57 | ···················· | ···················· | ····················· | ···················· | ···················· | ···················· | |
| L | 1842 hCoV-19/Switzerland/VD-ETHZ-100307/2020\|EPI_ISL_57 | ···················· | ···················· | ····················· | ···················· | ···················· | W··················· | |
| L | 1843 hCoV-19/Switzerland/VD-ETHZ-100324/2020\|EPI_ISL_57 | ···················· | ···················A | ····················· | ···················· | ···················· | ···················· | 1 |
| L | 1844 hCoV-19/Switzerland/VD-ETHZ-100327/2020\|EPI_ISL_57 | ···················· | ···················· | ····················· | ···················· | ···················· | ···················· | |
| L | 1845 hCoV-19/Switzerland/VD-ETHZ-100328/2020\|EPI_ISL_57 | ···················· | ·A·················· | ····················· | ···················· | ···················· | ···················· | |
| L | 1846 hCoV-19/Switzerland/VD-ETHZ-100329/2020\|EPI_ISL_57 | ···················· | ···················· | ····················· | ···················· | ···················· | ···················· | |
| L | 1847 hCoV-19/Switzerland/VD-ETHZ-100330/2020\|EPI_ISL_57 | ···················· | ·A·················· | ····················· | ···················· | ···················· | ···················· | |
| L | 1848 hCoV-19/Switzerland/VD-ETHZ-100331/2020\|EPI_ISL_57 | ···················· | ···················· | ····················· | ···················· | ···················· | ···················· | |
| L | 1849 hCoV-19/Switzerland/VD-ETHZ-100332/2020\|EPI_ISL_57 | ···················· | ···················· | ····················· | ···················· | ···················· | ···················· | |
| L | 1850 hCoV-19/Switzerland/VD-ETHZ-100333/2020\|EPI_ISL_57 | ···················· | ···················· | ····················· | ···················· | ···················· | ···················· | |
| L | 1851 hCoV-19/Switzerland/VD-ETHZ-100371/2020\|EPI_ISL_57 | ···················· | ···················· | ····················· | ···················· | ···················· | ···················· | |
| L | 1852 hCoV-19/Switzerland/ZH-ETHZ-100194/2020\|EPI_ISL_57 | ···················· | ···················· | ····················· | ···················· | ···················· | ···················· | |
| L | 1853 hCoV-19/Switzerland/ZH-ETHZ-100195/2020\|EPI_ISL_57 | ···················· | ···················· | ····················· | ···················· | ···················· | ···················· | |
| L | 1854 hCoV-19/Switzerland/ZH-ETHZ-100202/2020\|EPI_ISL_57 | ···················· | ···················· | ····················· | ···················· | ···················· | ···················· | |
| L | 1855 hCoV-19/USA/TX-DSHS-0645/2020\|EPI_ISL_576300\|2020- | ···················· | ···················· | ····················· | ···················· | ···················· | ···················· | |
| L | 1856 hCoV-19/Australia/NSW2829/2020\|EPI_ISL_577604\|2020 | ···················· | ···A················ | ····················· | ···················· | ···················· | ···················· | |
| L | 1857 hCoV-19/Czech Republic/NRL_6236/2020\|EPI_ISL_57762 | ···················· | ···················· | ····················· | ···················· | ···················· | ···················· | |
| L | 1858 hCoV-19/Czech Republic/NRL_7949/2020\|EPI_ISL_57762 | ···················· | ···················· | ····················· | ···················· | ···················· | ···················· | |
| L | 1859 hCoV-19/Czech Republic/NRL_7952/2020\|EPI_ISL_57762 | ···················· | ···················· | ····················· | ···················· | ···················· | ···················· | |
| L | 1860 hCoV-19/Czech Republic/NRL_7953/2020\|EPI_ISL_57762 | ···················· | ···················· | ····················· | ···················· | ···················· | ···················· | |
| L | 1861 hCoV-19/Czech Republic/NRL_8118-6/2020\|EPI_ISL_577 | ···················· | ···················· | ····················· | ···················· | ···················· | ···················· | |
| L | 1862 hCoV-19/Czech Republic/NRL_8118-7/2020\|EPI_ISL_577 | ···················· | ···················· | ····················· | ···················· | ···················· | ···················· | |
| L | 1863 hCoV-19/Czech Republic/NRL_6129/2020\|EPI_ISL_57762 | ···················· | ···················· | ····················· | ···················· | ···················· | ···················· | |
| L | 1864 hCoV-19/Czech Republic/NRL_6849-7/2020\|EPI_ISL_577 | ···················· | ···················· | ····················· | ···················· | ···················· | ···················· | |
| L | 1865 hCoV-19/Czech Republic/NRL_6849-10/2020\|EPI_ISL_57 | ···················· | ···················· | ····················· | ···················· | ···················· | ···················· | |
| L | 1866 hCoV-19/Czech Republic/NRL_6849-11/2020\|EPI_ISL_57 | ···················· | ···················· | ····················· | ···················· | ···················· | ···················· | |
| L | 1867 hCoV-19/Czech Republic/NRL_7164/2020\|EPI_ISL_57763 | ···················· | ···················· | ····················· | ···················· | ···················· | ···················· | |
| L | 1868 hCoV-19/Czech Republic/NRL_8458/2020\|EPI_ISL_57763 | ···················· | ···················· | ····················· | ···················· | ···················· | ···················· | |
| L | 1869 hCoV-19/Czech Republic/NRL_8459/2020\|EPI_ISL_57763 | ···················· | ···················· | ····················· | ···················· | ···················· | ···················· | |
| L | 1870 hCoV-19/Czech Republic/NRL_8460/2020\|EPI_ISL_57763 | ···················· | ···················· | ····················· | ···················· | ···················· | ···················· | |
| L | 1871 hCoV-19/Czech Republic/NRL_8461/2020\|EPI_ISL_57763 | ···················· | ···················· | ····················· | ···················· | ···················· | ···················· | |
| L | 1872 hCoV-19/Czech Republic/NRL_8496/2020\|EPI_ISL_57763 | ···················· | ···················· | ····················· | ···················· | ···················· | ···················· | |
| L | 1873 hCoV-19/India/MH-NIV-226874/2020\|EPI_ISL_577640\|20 | ···················· | ···················· | ····················· | ···················· | ···················· | ···················· | |
| L | 1874 hCoV-19/India/MH-NIV-212454/2020\|EPI_ISL_577643\|20 | ···················· | ···················· | ····················· | ···················· | ···················· | ···················· | |
| L | 1875 hCoV-19/India/MH-NIV-213875/2020\|EPI_ISL_577644\|20 | ···················· | ···················· | ····················· | ···················· | ···················· | ···················· | |
| L | 1876 hCoV-19/India/MH-NIV-215370/2020\|EPI_ISL_577645\|20 | ···················· | ···················· | ····················· | ···················· | ···················· | ···················· | |
| L | 1877 hCoV-19/India/MH-NIV-223127/2020\|EPI_ISL_577647\|20 | ···················· | ···················· | ····················· | ···················· | ···················· | ···················· | |
| L | 1878 hCoV-19/India/MH-NIV-210557/2020\|EPI_ISL_577648\|20 | ···················· | ···················· | ····················· | ···················· | ···················· | ···················· | |
| L | 1879 hCoV-19/India/MH-NIV-223537/2020\|EPI_ISL_577649\|20 | ···················· | ···················· | ····················· | ···················· | ···················· | ···················· | |
| L | 1880 hCoV-19/India/MH-NIV-221624/2020\|EPI_ISL_577651\|20 | ···················· | ···················· | ····················· | ···················· | ···················· | ···················· | |
| L | 1881 hCoV-19/India/MH-NIV-222131/2020\|EPI_ISL_577652\|20 | ···················· | ···················· | ····················· | ···················· | ···················· | ···················· | |

| | | | | | | | | |
|---|---|---|---|---|---|---|---|---|
| L | 1882 | hCoV-19/India/MH-NIV-220301/2020\|EPI_ISL_577655\|20 | ···················· | ···················· | ···················· | ···················· | ···················· | ···················· |
| L | 1883 | hCoV-19/India/MH-NIV-223126/2020\|EPI_ISL_577658\|20 | ···················· | ···················· | ···················· | ···················· | ···················· | ···················· |
| L | 1884 | hCoV-19/India/MH-NIV-216133/2020\|EPI_ISL_577659\|20 | ···················· | ···················· | ···················· | ···················· | ···················· | ···················· |
| L | 1885 | hCoV-19/India/MH-NIV-216500/2020\|EPI_ISL_577660\|20 | ···················· | ···················· | ···················· | ···················· | ···················· | ···················· |
| L | 1886 | hCoV-19/India/MH-NIV-207705/2020\|EPI_ISL_577661\|20 | ···················· | ·A·················· | ···················· | ···················· | ···················· | ···A················ |
| L | 1887 | hCoV-19/India/MH-NIV-216641/2020\|EPI_ISL_577662\|20 | ···················· | ···················· | ···················· | ···················· | ···················· | ···················· |
| L | 1888 | hCoV-19/India/MH-NIV-209697/2020\|EPI_ISL_577663\|20 | ···················· | ···················· | ···················· | ···················· | ···················· | ···················· |
| L | 1889 | hCoV-19/India/MH-NIV-216244/2020\|EPI_ISL_577664\|20 | ···················· | ···················· | ···················· | ···················· | ···················· | ···················· |
| L | 1890 | hCoV-19/India/MH-NIV-216880/2020\|EPI_ISL_577665\|20 | ···················· | ···················· | ···················· | ···················· | ···················· | ···················· |
| L | 1891 | hCoV-19/India/MH-NIV-210934/2020\|EPI_ISL_577670\|20 | ···················· | ···················· | ···················· | ···················· | ···················· | ···················· |
| L | 1892 | hCoV-19/India/MH-NIV-197096/2020\|EPI_ISL_577672\|20 | ···················· | ·A·················· | ···················· | ···················· | ···················· | ···A················ |
| L | 1894 | hCoV-19/India/MH-NIV-229804/2020\|EPI_ISL_577677\|20 | ···················· | ···················· | ···················· | ···················· | ···················· | ···················· |
| L | 1895 | hCoV-19/USA/LA-EVTL503/2020\|EPI_ISL_578907\|2020-06 | ···················· | ···················· | ···················· | ···················· | ···················· | ···················· |
| L | 1896 | hCoV-19/USA/LA-EVTL506/2020\|EPI_ISL_578910\|2020-06 | ···················· | ···················· | ···················· | ···················· | ···················· | ···················· |
| L | 1897 | hCoV-19/USA/LA-EVTL507/2020\|EPI_ISL_578911\|2020-06 | ···················· | ···················· | ···················· | ···················· | ···················· | ···················· |
| L | 1898 | hCoV-19/USA/LA-EVTL508/2020\|EPI_ISL_578912\|2020-07 | ···················· | ···················· | ···················· | ···················· | ···················· | ···················· |
| L | 1899 | hCoV-19/USA/LA-EVTL509/2020\|EPI_ISL_578913\|2020-07 | ···················· | ···················· | ···················· | ···················· | ···················· | ···················· |
| L | 1900 | hCoV-19/USA/LA-EVTL510/2020\|EPI_ISL_578914\|2020-06 | ···················· | ···················· | ···················· | ···················· | ···················· | ···················· |
| L | 1901 | hCoV-19/USA/LA-EVTL511/2020\|EPI_ISL_578915\|2020-06 | ···················· | ···················· | ···················· | ···················· | ···················· | ···················· |
| L | 1902 | hCoV-19/USA/LA-EVTL512/2020\|EPI_ISL_578916\|2020-06 | ···················· | ·············A······ | ···················· | ···················· | ···················· | ···················· |
| L | 1903 | hCoV-19/USA/LA-EVTL513/2020\|EPI_ISL_578917\|2020-06 | ···················· | ···················· | ···················· | ···················· | ···················· | ···················· |
| L | 1904 | hCoV-19/USA/LA-EVTL514/2020\|EPI_ISL_578918\|2020-06 | ···················· | ···················· | ···················· | ···················· | ···················· | ···················· |
| L | 1905 | hCoV-19/USA/LA-EVTL515/2020\|EPI_ISL_578919\|2020-08 | ···················· | ···················· | ···················· | ···················· | ···················· | ···················· |
| L | 1906 | hCoV-19/USA/LA-EVTL516/2020\|EPI_ISL_578920\|2020-06 | ···················· | ···················· | ···················· | ···················· | ·····C·············· | ···················· |
| L | 1907 | hCoV-19/USA/LA-EVTL517/2020\|EPI_ISL_578921\|2020-06 | ···················· | ···················· | ···················· | ···················· | ···················· | ···················· |
| L | 1908 | hCoV-19/USA/LA-EVTL518/2020\|EPI_ISL_578922\|2020-06 | ···················· | ···················· | ···················· | ···················· | ···················· | ···················· |
| L | 1909 | hCoV-19/USA/LA-EVTL519/2020\|EPI_ISL_578923\|2020-08 | ···················· | ···················· | ···················· | ···················· | ···················· | ···················· |
| L | 1910 | hCoV-19/USA/LA-EVTL520/2020\|EPI_ISL_578924\|2020-06 | ···················· | ···················· | ···················· | ···················· | ···················· | ···················· |
| L | 1911 | hCoV-19/USA/LA-EVTL521/2020\|EPI_ISL_578925\|2020-06 | ···················· | ···················· | ···················· | ···················· | ···················· | ···················· |
| L | 1912 | hCoV-19/USA/LA-EVTL522/2020\|EPI_ISL_578926\|2020-06 | ···················· | ···················· | ···················· | ···················· | ·····C·············· | ···················· |
| L | 1913 | hCoV-19/USA/LA-EVTL523/2020\|EPI_ISL_578927\|2020-08 | ···················· | ···················· | ···················· | ···················· | ···················· | ···················· |
| L | 1914 | hCoV-19/USA/LA-EVTL524/2020\|EPI_ISL_578928\|2020-08 | ···················· | ·············A······ | ···················· | ···················· | ···················· | ···················· |
| L | 1915 | hCoV-19/USA/LA-EVTL525/2020\|EPI_ISL_578929\|2020-08 | ···················· | ·A·················· | ···················· | ···················· | ···················· | ···················· |
| L | 1916 | hCoV-19/USA/LA-EVTL526/2020\|EPI_ISL_578930\|2020-08 | ···················· | ·A·················· | ···················· | ···················· | ···················· | ···················· |
| L | 1917 | hCoV-19/USA/LA-EVTL527/2020\|EPI_ISL_578931\|2020-06 | ···················· | ···················· | ···················· | ···················· | ···················· | ···················· |
| L | 1918 | hCoV-19/USA/LA-EVTL530/2020\|EPI_ISL_578934\|2020-08 | ···················· | ···················· | ···················· | ···················· | ···················· | ···················· |
| L | 1919 | hCoV-19/USA/LA-EVTL531/2020\|EPI_ISL_578935\|2020-08 | ···················· | ···················· | ···················· | ···················· | ···················· | ···················· |
| L | 1920 | hCoV-19/USA/LA-EVTL539/2020\|EPI_ISL_578943\|2020-06 | ···················· | ···················· | ···················· | ···················· | ···················· | ···················· |
| L | 1921 | hCoV-19/USA/LA-EVTL540/2020\|EPI_ISL_578944\|2020-06 | ···················· | ···················· | ···················· | ···················· | ···················· | ···················· |
| L | 1922 | hCoV-19/USA/LA-EVTL541/2020\|EPI_ISL_578945\|2020-06 | ···················· | ···················· | ···················· | ···················· | ···················· | ···················· |
| L | 1923 | hCoV-19/USA/LA-EVTL542/2020\|EPI_ISL_578946\|2020-06 | ···················· | ···················· | ···················· | ···················· | ···················· | ···················· |
| L | 1924 | hCoV-19/USA/LA-EVTL543/2020\|EPI_ISL_578947\|2020-06 | ···················· | ·A·················· | ···················· | ···················· | ···················· | ···················· |
| L | 1925 | hCoV-19/USA/LA-EVTL544/2020\|EPI_ISL_578948\|2020-06 | ···················· | ···················· | ···················· | ···················· | ···················· | ···················· |
| L | 1926 | hCoV-19/USA/LA-EVTL545/2020\|EPI_ISL_578949\|2020-08 | ···················· | ···················· | ···················· | ···················· | ···················· | ···················· |
| L | 1927 | hCoV-19/USA/LA-EVTL546/2020\|EPI_ISL_578950\|2020-08 | ···················· | ···················· | ···················· | ···················· | ···················· | ···················· |
| L | 1928 | hCoV-19/USA/LA-EVTL547/2020\|EPI_ISL_578951\|2020-08 | ···················· | ···················· | ···················· | ···················· | ···················· | ···················· |
| L | 1929 | hCoV-19/USA/LA-EVTL548/2020\|EPI_ISL_578952\|2020-06 | ···················· | ···················· | ···················· | ···················· | ···················· | ···················· |
| L | 1930 | hCoV-19/USA/LA-EVTL549/2020\|EPI_ISL_578953\|2020-08 | ···················· | ···················· | ···················· | ···················· | ···················· | ···················· |
| L | 1931 | hCoV-19/USA/LA-EVTL550/2020\|EPI_ISL_578954\|2020-06 | ···················· | ···················· | ···················· | ···················· | ···················· | ···················· |
| L | 1932 | hCoV-19/USA/LA-EVTL551/2020\|EPI_ISL_578955\|2020-08 | ···················· | ···················· | ···················· | ···················· | ···················· | ···················· |
| L | 1933 | hCoV-19/USA/LA-EVTL552/2020\|EPI_ISL_578956\|2020-08 | ···················· | ···················· | ···················· | ···················· | ···················· | ···················· |
| L | 1934 | hCoV-19/USA/LA-EVTL553/2020\|EPI_ISL_578957\|2020-08 | ···················· | ···················· | ···················· | ···················· | ···················· | ···················· |
| L | 1935 | hCoV-19/USA/LA-EVTL554/2020\|EPI_ISL_578958\|2020-08 | ···················· | ···················· | ···················· | ···················· | ···················· | ···················· |
| L | 1936 | hCoV-19/USA/LA-EVTL555/2020\|EPI_ISL_578959\|2020-08 | ···················· | ···················· | ···················· | ···················· | ···················· | ···················· |
| L | 1937 | hCoV-19/USA/LA-EVTL556/2020\|EPI_ISL_578960\|2020-08 | ···················· | ···················· | ···················· | ···················· | ···················· | ···················· |
| L | 1938 | hCoV-19/USA/LA-EVTL557/2020\|EPI_ISL_578961\|2020-08 | ···················· | ···················· | ···················· | ···················· | ···················· | ···················· |
| L | 1939 | hCoV-19/USA/LA-EVTL558/2020\|EPI_ISL_578962\|2020-08 | ···················· | ···················· | ···················· | ···················· | ···················· | ···················· |
| L | 1940 | hCoV-19/USA/LA-EVTL559/2020\|EPI_ISL_578963\|2020-08 | ···················· | ···················· | ···················· | ···················· | ···················· | ···················· |
| L | 1941 | hCoV-19/USA/LA-EVTL560/2020\|EPI_ISL_578964\|2020-08 | ···················· | ···················· | ···················· | ···················· | ···················· | ···················· |
| L | 1942 | hCoV-19/USA/LA-EVTL561/2020\|EPI_ISL_578965\|2020-08 | ···················· | ···················· | ···················· | ···················· | ···················· | ···················· |
| L | 1943 | hCoV-19/USA/LA-EVTL562/2020\|EPI_ISL_578966\|2020-08 | ···················· | ···················· | ···················· | ···················· | ···················· | ···················· |
| L | 1944 | hCoV-19/USA/LA-EVTL563/2020\|EPI_ISL_578967\|2020-06 | ···················· | ···················· | ···················· | ···················· | ···················· | ···················· |
| L | 1945 | hCoV-19/USA/LA-EVTL564/2020\|EPI_ISL_578968\|2020-06 | ···················· | ···················· | ···················· | ···················· | ···················· | ···················· |
| L | 1946 | hCoV-19/USA/LA-EVTL565/2020\|EPI_ISL_578969\|2020-06 | ···················· | ···················· | ···················· | ···················· | ···················· | ···················· |
| L | 1947 | hCoV-19/USA/LA-EVTL566/2020\|EPI_ISL_578970\|2020-06 | ···················· | ···················· | ···················· | ···················· | ···················· | ···················· |
| L | 1948 | hCoV-19/USA/LA-EVTL567/2020\|EPI_ISL_578971\|2020-06 | ···················· | ···················· | ···················· | ···················· | ···················· | ···················· |
| L | 1949 | hCoV-19/USA/LA-EVTL568/2020\|EPI_ISL_578972\|2020-06 | ···················· | ···················· | ···················· | ···················· | ···················· | ···················· |
| L | 1950 | hCoV-19/USA/LA-EVTL569/2020\|EPI_ISL_578973\|2020-06 | ···················· | ···················· | ···················· | ···················· | ···················· | ···················· |
| L | 1951 | hCoV-19/USA/LA-EVTL570/2020\|EPI_ISL_578974\|2020-06 | ···················· | ···················· | ···················· | ···················· | ···················· | ···················· |
| L | 1952 | hCoV-19/USA/LA-EVTL571/2020\|EPI_ISL_578975\|2020-06 | ···················· | ···················· | ···················· | ···················· | ···················· | ···················· |
| L | 1953 | hCoV-19/USA/LA-EVTL572/2020\|EPI_ISL_578976\|2020-06 | ···················· | ···················· | ···················· | ···················· | ···················· | ···················· |
| L | 1954 | hCoV-19/USA/LA-EVTL573/2020\|EPI_ISL_578977\|2020-06 | ···················· | ···················· | ···················· | ···················· | ···················· | ···················· |
| L | 1955 | hCoV-19/USA/LA-EVTL574/2020\|EPI_ISL_578978\|2020-06 | ···················· | ···················· | ···················· | ···················· | ···················· | ···················· |
| L | 1956 | hCoV-19/USA/LA-EVTL575/2020\|EPI_ISL_578979\|2020-06 | ···················· | ···················· | ···················· | ···················· | ···················· | ···················· |
| L | 1957 | hCoV-19/USA/LA-EVTL576/2020\|EPI_ISL_578980\|2020-06 | ···················· | ···················· | ···················· | ···················· | ···················· | ···················· |
| L | 1958 | hCoV-19/USA/LA-EVTL577/2020\|EPI_ISL_578981\|2020-06 | ···················· | ···················· | ···················· | ···················· | ···················· | ···················· |
| L | 1959 | hCoV-19/USA/LA-EVTL578/2020\|EPI_ISL_578982\|2020-06 | ···················· | ···················· | ···················· | ···················· | ···················· | ···················· |
| L | 1960 | hCoV-19/USA/LA-EVTL579/2020\|EPI_ISL_578983\|2020-06 | ···················· | ···················· | ···················· | ···················· | ···················· | ···················· |
| L | 1961 | hCoV-19/USA/LA-EVTL580/2020\|EPI_ISL_578984\|2020-06 | ···················· | ···················· | ···················· | ···················· | ···················· | ···················· |
| L | 1962 | hCoV-19/USA/LA-EVTL581/2020\|EPI_ISL_578985\|2020-06 | ···················· | ···················· | ···················· | ···················· | ···················· | ···················· |

L 1963 hCoV-19/USA/LA-EVTL582/2020|EPI_ISL_578986|2020-06 .................... .................... .................... .................... .................... ....................
L 1964 hCoV-19/USA/LA-EVTL583/2020|EPI_ISL_578987|2020-06 .................... .................... .................... .................... .................... ....................
L 1965 hCoV-19/USA/LA-EVTL584/2020|EPI_ISL_578988|2020-06 .................... .................... .................... .................... .................... ....................
L 1966 hCoV-19/USA/LA-EVTL585/2020|EPI_ISL_578989|2020-06 .................... .................... .................... .................... .................... ....................
L 1967 hCoV-19/USA/LA-EVTL586/2020|EPI_ISL_578990|2020-06 .................... .................... .................... .................... .................... ....................
L 1968 hCoV-19/USA/LA-EVTL587/2020|EPI_ISL_578991|2020-06 .................... .................... .................... .................... .................... ....................
L 1969 hCoV-19/USA/LA-EVTL588/2020|EPI_ISL_578992|2020-06 .................... .................... .................... .................... .................... ....................
L 1970 hCoV-19/USA/LA-EVTL589/2020|EPI_ISL_578993|2020-06 .................... .................... .................... .................... .................... ....................
L 1971 hCoV-19/USA/LA-EVTL590/2020|EPI_ISL_578994|2020-06 .................... .................... .................... .................... .................... ....................
L 1972 hCoV-19/USA/LA-EVTL591/2020|EPI_ISL_578995|2020-06 .................... .................... .................... .................... .................... ....................
L 1973 hCoV-19/USA/LA-EVTL592/2020|EPI_ISL_578996|2020-06 .................... .................... .................... .................... .................... ....................
L 1974 hCoV-19/USA/LA-EVTL593/2020|EPI_ISL_578997|2020-06 .................... .................... .................... .................... .................... ....................
L 1975 hCoV-19/USA/LA-EVTL594/2020|EPI_ISL_578998|2020-06 .................... .................... .................... .................... .................... ....................
L 1976 hCoV-19/USA/LA-EVTL595/2020|EPI_ISL_578999|2020-06 .................... .................... .................... .................... .................... ....................
L 1977 hCoV-19/Spain/CB-IBV-98007068/2020|EPI_ISL_582100| .................... .................... .................... .................... .................... ....................
L 1978 hCoV-19/Spain/CB-IBV-98007069/2020|EPI_ISL_582101| .................... .................... .................... .................... .................... ....................
L 1979 hCoV-19/Spain/CB-IBV-98007070/2020|EPI_ISL_582102| .................... .................... .................... .................... .................... ....................
L 1980 hCoV-19/Spain/CB-IBV-98007073/2020|EPI_ISL_582103| .................... .................... .................... .................... .................... ....................
L 1982 hCoV-19/Spain/CB-IBV-98007075/2020|EPI_ISL_582105| .................... .................... .................... .................... .................... ....................
L 1983 hCoV-19/Spain/CB-IBV-98007076/2020|EPI_ISL_582106| .................... .................... .................... .................... .................... ....................
L 1984 hCoV-19/Spain/CB-IBV-98007079/2020|EPI_ISL_582107| .................... .................... .................... .................... .................... ....................
L 1985 hCoV-19/Spain/CB-IBV-98007080/2020|EPI_ISL_582108| .................... .................... .................... .................... .................... ....................
L 1986 hCoV-19/Spain/CB-IBV-98007086/2020|EPI_ISL_582109| .................... .................... .................... .................... .................... ....................
L 1987 hCoV-19/France/ARA-CDP82203/2020|EPI_ISL_582110|20 .................... .................... .................... .................... .................... ....................
L 1988 hCoV-19/France/ARA-CDP81878/2020|EPI_ISL_582111|20 .................... .................... .................... .................... .................... ....................
L 1989 hCoV-19/France/ARA-CDP29560/2020|EPI_ISL_582112|20 .................... .................... .................... .................... .................... ....................
L 1990 hCoV-19/France/ARA-CDP20536/2020|EPI_ISL_582114|20 .................... .................... .................... .................... .................... ....................
L 1991 hCoV-19/France/ARA-CDP29592/2020|EPI_ISL_582115|20 .................... .................... .................... .................... .................... ....................
L 1992 hCoV-19/France/ARA-CDP81800/2020|EPI_ISL_582116|20 .................... .................... .................... .................... .................... ....................
L 1993 hCoV-19/France/ARA-CDP82238/2020|EPI_ISL_582118|20 .................... .................... .................... .................... .................... ....................
L 1994 hCoV-19/France/ARA-CDP82109/2020|EPI_ISL_582119|20 .................... .................... .................... .................... .................... ....................
L 1995 hCoV-19/France/ARA-CDP78128/2020|EPI_ISL_582120|20 .................... .................... .................... .................... .................... ....................
L 1996 hCoV-19/France/ARA-59384/2020|EPI_ISL_582121|2020- .................... .................... .................... .................... .................... ....................
L 1997 hCoV-19/France/ARA-71991/2020|EPI_ISL_582122|2020- .................... .................... .................... .................... .................... ....................
L 1998 hCoV-19/Italy/TUS-DBM-1/2020|EPI_ISL_582123|2020-0 .................... .................... .................... .................... .................... ....................
L 1999 hCoV-19/Malaysia/MGI-MAEPS41/2020|EPI_ISL_582124|2 .................... .................... .................... .................... .................... ....................
L 2000 hCoV-19/United Arab Emirates/skmc-920168117/2020|E .................... .................... .................... .................... .................... ....................
L 2001 hCoV-19/United Arab Emirates/skmc-920569110/2020|E .................... .................... .................... .................... .................... ....................
L 2002 hCoV-19/Belgium/ITM-C5182/2020|EPI_ISL_582127|2020 .................... .................... .................... .................... .................... ....................
L 2003 hCoV-19/Belgium/ITM-C5183/2020|EPI_ISL_582128|2020 .................... .................... .................... .................... .................... ....................
L 2004 hCoV-19/Belgium/ITM-C5184/2020|EPI_ISL_582129|2020 .................... .................... .................... .................... .................... ....................
L 2005 hCoV-19/Belgium/ITM-C5185/2020|EPI_ISL_582130|2020 .................... .................... .................... .................... .................... ....................
L 2006 hCoV-19/Belgium/ITM-C5186/2020|EPI_ISL_582131|2020 .................... .................... .................... .................... .................... ....................
L 2007 hCoV-19/Belgium/ITM-C5188/2020|EPI_ISL_582132|2020 .................... .................... .................... .................... .................... ....................
L 2008 hCoV-19/USA/NY-MSK-3A/2020|EPI_ISL_583431|2020-03- .................... .................... .................... .................... .................... ....................
L 2009 hCoV-19/USA/NY-MSK-3B/2020|EPI_ISL_583432|2020-04- .................... .................... .................... .................... .................... ....................
L 2010 hCoV-19/USA/NY-MSK-3C/2020|EPI_ISL_583433|2020-04- .................... .................... .................... .................... .................... ....................
L 2011 hCoV-19/USA/NY-MSK-3Ai/2020|EPI_ISL_583434|2020-03 .................... .................... .................... .................... .................... ....................
L 2012 hCoV-19/USA/NY-MSK-3Bi/2020|EPI_ISL_583435|2020-04 .................... .................... .................... .................... .................... ....................
L 2013 hCoV-19/USA/NY-MSK-3Ci/2020|EPI_ISL_583436|2020-04 .................... .................... .................... .................... .................... ....................
L 2014 hCoV-19/USA/NY-MSK-3D/2020|EPI_ISL_583437|2020-04- .................... .................... .................... .................... .................... ....................
L 2015 hCoV-19/USA/NY-MSK-4A/2020|EPI_ISL_583439|2020-04- .................... .................... .................... .................... .................... ....................
L 2016 hCoV-19/USA/NY-MSK-4C/2020|EPI_ISL_583440|2020-05- .................... .................... .................... .................... .................... ....................
L 2017 hCoV-19/USA/NY-MSK-4Ai/2020|EPI_ISL_583441|2020-04 .................... .................... .................... .................... .................... ....................
L 2018 hCoV-19/USA/NY-MSK-4Ci/2020|EPI_ISL_583442|2020-05 .................... .................... .................... .................... .................... ....................
L 2019 hCoV-19/USA/NY-MSK-4E/2020|EPI_ISL_583443|2020-05- .................... .................... .................... .................... .................... ....................
L 2020 hCoV-19/USA/NY-MSK-5B/2020|EPI_ISL_583445|2020-04- .................... .................... .................... .................... .................... ....................
L 2021 hCoV-19/USA/NY-MSK-6C/2020|EPI_ISL_583446|2020-04- .................... .................... .................... .................... .................... ....................
L 2022 hCoV-19/USA/NY-MSK-6E/2020|EPI_ISL_583448|2020-05- .................... .................... .................... .................... .................... ....................
L 2023 hCoV-19/USA/NY-MSK-6F/2020|EPI_ISL_583449|2020-05- .................... .................... .................... .................... .................... ....................
L 2024 hCoV-19/USA/NY-MSK-8A/2020|EPI_ISL_583456|2020-04- .................... .................... .................... .................... .................... ....................
L 2025 hCoV-19/USA/NY-MSK-8B/2020|EPI_ISL_583457|2020-05- .................... .................... .................... .................... .................... ....................
L 2026 hCoV-19/USA/NY-MSK-8Ai/2020|EPI_ISL_583458|2020-04 .................... .................... .................... .................... .................... ....................
L 2027 hCoV-19/USA/NY-MSK-8Bi/2020|EPI_ISL_583459|2020-05 .................... ...................A .................... .................... .................... .................... 1
L 2028 hCoV-19/USA/NY-MSK-8C/2020|EPI_ISL_583460|2020-05- .................... .................... .................... .................... .................... ....................
L 2029 hCoV-19/USA/NY-MSK-9A/2020|EPI_ISL_583461|2020-03- .................... .................... .................... .................... .................... ....................
L 2030 hCoV-19/USA/NY-MSK-9B/2020|EPI_ISL_583462|2020-03- .................... .................... .................... .................... .................... ....................
L 2031 hCoV-19/USA/NY-MSK-9Ai/2020|EPI_ISL_583463|2020-03 .................... .................... .................... .................... .................... ....................
L 2032 hCoV-19/USA/NY-MSK-9Bi/2020|EPI_ISL_583464|2020-03 .................... .................... .................... .................... .................... ....................
L 2033 hCoV-19/USA/NY-MSK-10B/2020|EPI_ISL_583465|2020-05 .................... .................... .................... .................... .................... ....................
L 2034 hCoV-19/USA/NY-MSK-11A/2020|EPI_ISL_583466|2020-03 .................... .................... .................... .................... .................... ....................
L 2035 hCoV-19/USA/NY-MSK-11Ai/2020|EPI_ISL_583467|2020-0 .................... .................... .................... .................... .................... ....................
L 2036 hCoV-19/USA/NY-MSK-11D/2020|EPI_ISL_583468|2020-04 .................... .................... .................... .................... .................... ....................
L 2037 hCoV-19/USA/NY-MSK-13A/2020|EPI_ISL_583469|2020-03 .................... .................... .................... .................... .................... ....................
L 2038 hCoV-19/USA/NY-MSK-13B/2020|EPI_ISL_583470|2020-05 .................... .................... .................... .................... .................... ....................
L 2039 hCoV-19/USA/NY-MSK-14A/2020|EPI_ISL_583472|2020-03 .................... .................... .................... .................... .................... ....................
L 2040 hCoV-19/USA/NY-MSK-14Ai/2020|EPI_ISL_583473|2020-0 .................... .................... .................... .................... .................... ....................
L 2041 hCoV-19/USA/NY-MSK-16A/2020|EPI_ISL_583475|2020-04 .................... .................... .................... .................... .................... ....................
L 2042 hCoV-19/USA/NY-MSK-16B/2020|EPI_ISL_583476|2020-06 .................... .................... .................... .................... .................... ....................
L 2043 hCoV-19/USA/NY-MSK-16Ai/2020|EPI_ISL_583477|2020-0 .................... .................... .................... .................... .................... ....................

| | | | | | | | | |
|---|---|---|---|---|---|---|---|---|
| L | 2044 | hCoV-19/USA/NY-MSK-17B/2020\|EPI_ISL_583478\|2020-05 | ···················· | ···················· | ···················· | ···················· | ···················· | ···················· |
| L | 2045 | hCoV-19/USA/NY-MSK-20A/2020\|EPI_ISL_583479\|2020-03 | ···················· | ···················· | ···················· | ···················· | ···················· | ···················· |
| L | 2046 | hCoV-19/USA/NY-MSK-20E/2020\|EPI_ISL_583480\|2020-05 | ···················· | ···················· | ···················· | ···················· | ···················· | ···················· |
| L | 2047 | hCoV-19/Australia/VIC13276/2020\|EPI_ISL_594144\|202 | ···················· | ···················· | ···················· | ···················· | ···················· | ···················· |
| L | 2048 | hCoV-19/Australia/VIC13568/2020\|EPI_ISL_594147\|202 | ···················· | ···················· | ···················· | ···················· | ···················· | ···················· |
| L | 2049 | hCoV-19/England/ALDP-9BCF58/2020\|EPI_ISL_573083\|20 | ···················· | ···················· | ···················· | ···················· | ···················· | ···················· |
| L | 2050 | hCoV-19/England/ALDP-9BCF67/2020\|EPI_ISL_573084\|20 | ···················· | ·A·················· | ···················· | ···················· | ···················· | ···A················ |
| L | 2054 | hCoV-19/England/ALDP-9CC1AA/2020\|EPI_ISL_573089\|20 | ···················· | ···················· | ···················· | ···················· | ···················· | ···················· |
| L | 2057 | hCoV-19/England/ALDP-9CC201/2020\|EPI_ISL_573092\|20 | ···················· | ···················· | ···················· | ···················· | ···················· | ···················· |
| L | 2061 | hCoV-19/England/ALDP-9CC2F2/2020\|EPI_ISL_573096\|20 | ···················· | ···················· | ···················· | ···················· | ···················· | ···················· |
| L | 2063 | hCoV-19/England/ALDP-9CC31D/2020\|EPI_ISL_573098\|20 | ···················· | ···················· | ···················· | ···················· | ···················· | ···················· |
| L | 2065 | hCoV-19/USA/LA-EVTL496/2020\|EPI_ISL_578900\|2020-07 | ···················· | ···················· | ···················· | ···················· | ···················· | ···················· |
| L | 2066 | hCoV-19/USA/LA-EVTL497/2020\|EPI_ISL_578901\|2020-07 | ···················· | ···················· | ···················· | ···················· | ···················· | ···················· |
| L | 2067 | hCoV-19/USA/LA-EVTL498/2020\|EPI_ISL_578902\|2020-07 | ···················· | ···················· | ···················· | ···················· | ···················· | ···················· |
| L | 2068 | hCoV-19/USA/LA-EVTL499/2020\|EPI_ISL_578903\|2020-07 | ···················· | ···················· | ···················· | ···················· | ···················· | ···················· |
| L | 2069 | hCoV-19/USA/LA-EVTL500/2020\|EPI_ISL_578904\|2020-07 | ···················· | ···················· | ···················· | ···················· | ···················· | ···················· |
| L | 2070 | hCoV-19/USA/LA-EVTL501/2020\|EPI_ISL_578905\|2020-07 | ···················· | ···················· | ···················· | ···················· | ···················· | ···················· |
| L | 2071 | hCoV-19/USA/LA-EVTL502/2020\|EPI_ISL_578906\|2020-07 | ···················· | ···················· | ···················· | ···················· | ···················· | ···················· |
| L | 2072 | hCoV-19/USA/LA-EVTL504/2020\|EPI_ISL_578908\|2020-06 | ···················· | ···················· | ···················· | ···················· | ···················· | ···················· |
| L | 2073 | hCoV-19/USA/LA-EVTL505/2020\|EPI_ISL_578909\|2020-06 | ···················· | ···················· | ···················· | ···················· | ···················· | ···················· |
| L | 2078 | hCoV-19/New Zealand/20VR2091/2020\|EPI_ISL_579227\|2 | ···················· | ···················· | ···················· | ···················· | ···················· | ···················· |
| L | 2080 | hCoV-19/New Zealand/20VR2101/2020\|EPI_ISL_579229\|2 | ···················· | ···················· | ···················· | ···················· | ···················· | ···················· |
| L | 2081 | hCoV-19/New Zealand/20VR2103/2020\|EPI_ISL_579230\|2 | ···················· | ···················· | ···················· | ···················· | ···················· | ···················· |
| L | 2082 | hCoV-19/New Zealand/20VR2471/2020\|EPI_ISL_579233\|2 | ···················· | ···················· | ···················· | ···················· | ···················· | ···················· |
| L | 2083 | hCoV-19/New Zealand/20VR2474/2020\|EPI_ISL_579236\|2 | ···················· | ···················· | ···················· | ···················· | ···················· | ···················· |
| L | 2084 | hCoV-19/New Zealand/20VR2476/2020\|EPI_ISL_579237\|2 | ···················· | ···················· | ···················· | ···················· | ···················· | ···················· |
| L | 2086 | hCoV-19/New Zealand/20VR2479/2020\|EPI_ISL_579240\|2 | ···················· | ···················· | ···················· | ···················· | ···················· | ···················· |
| L | 2087 | hCoV-19/New Zealand/20VR2486/2020\|EPI_ISL_579243\|2 | ···················· | ···················· | ···················· | ···················· | ···················· | ···················· |
| L | 2088 | hCoV-19/New Zealand/20VR2488/2020\|EPI_ISL_579245\|2 | ···················· | ···················· | ···················· | ···················· | ···················· | ···················· |
| L | 2089 | hCoV-19/New Zealand/20VR2489/2020\|EPI_ISL_579246\|2 | ···················· | ···················· | ···················· | ···················· | ···················· | ···················· |
| L | 2090 | hCoV-19/New Zealand/20VR2490/2020\|EPI_ISL_579247\|2 | ···················· | ···················· | ···················· | ···················· | ···················· | ···················· |
| L | 2091 | hCoV-19/New Zealand/20VR2491/2020\|EPI_ISL_579248\|2 | ···················· | ···················· | ···················· | ···················· | ···················· | ···················· |
| L | 2092 | hCoV-19/New Zealand/20VR2492/2020\|EPI_ISL_579249\|2 | ···················· | ···················· | ···················· | ···················· | ···················· | ···················· |
| L | 2093 | hCoV-19/New Zealand/20VR2494/2020\|EPI_ISL_579251\|2 | ···················· | ···················· | ···················· | ···················· | ···················· | ···················· |
| L | 2094 | hCoV-19/New Zealand/20VR2497/2020\|EPI_ISL_579252\|2 | ···················· | ···················· | ···················· | ···················· | ···················· | ···················· |
| L | 2095 | hCoV-19/New Zealand/20VR2498/2020\|EPI_ISL_579253\|2 | ···················· | ···················· | ···················· | ···················· | ···················· | ···················· |
| L | 2097 | hCoV-19/New Zealand/20VR2501/2020\|EPI_ISL_579256\|2 | ···················· | ···················· | ···················· | ···················· | ···················· | ···················· |
| L | 2099 | hCoV-19/New Zealand/20VR2507/2020\|EPI_ISL_579261\|2 | ···················· | ···················· | ···················· | ···················· | ···················· | ···················· |
| L | 2101 | hCoV-19/New Zealand/20VR2517/2020\|EPI_ISL_579263\|2 | ···················· | ···················· | ···················· | ···················· | ···················· | ···················· |
| L | 2103 | hCoV-19/New Zealand/20VR2519/2020\|EPI_ISL_579265\|2 | ···················· | ···················· | ···················· | ···················· | ···················· | ···················· |
| L | 2104 | hCoV-19/New Zealand/20VR2520/2020\|EPI_ISL_579266\|2 | ···················· | ···················· | ···················· | ···················· | ···················· | ···················· |
| L | 2109 | hCoV-19/New Zealand/20VR2526/2020\|EPI_ISL_579271\|2 | ···················· | ···················· | ···················· | ···················· | ···················· | ···················· |
| L | 2110 | hCoV-19/New Zealand/20VR2527/2020\|EPI_ISL_579272\|2 | ···················· | ···················· | ···················· | ···················· | ···················· | ···················· |
| L | 2113 | hCoV-19/New Zealand/20VR2532/2020\|EPI_ISL_579277\|2 | ···················· | ···················· | ···················· | ···················· | ···················· | ···················· |
| L | 2114 | hCoV-19/New Zealand/20VR2533/2020\|EPI_ISL_579278\|2 | ···················· | ···················· | ···················· | ···················· | ···················· | ···················· |
| L | 2116 | hCoV-19/New Zealand/20VR2535/2020\|EPI_ISL_579280\|2 | ···················· | ···················· | ···················· | ···················· | ···················· | ···················· |
| L | 2119 | hCoV-19/New Zealand/20VR2538/2020\|EPI_ISL_579283\|2 | ···················· | ···················· | ···················· | ···················· | ···················· | ···················· |
| L | 2122 | hCoV-19/New Zealand/20VR2545/2020\|EPI_ISL_579288\|2 | ···················· | ···················· | ···················· | ···················· | ···················· | ···················· |
| L | 2123 | hCoV-19/New Zealand/20VR2546/2020\|EPI_ISL_579289\|2 | ···················· | ···················· | ···················· | ···················· | ···················· | ···················· |
| L | 2124 | hCoV-19/New Zealand/20VR2547/2020\|EPI_ISL_579290\|2 | ···················· | ···················· | ···················· | ···················· | ···················· | ···················· |
| L | 2125 | hCoV-19/New Zealand/20VR2548/2020\|EPI_ISL_579291\|2 | ···················· | ···················· | ···················· | ···················· | ···················· | ···················· |
| L | 2126 | hCoV-19/New Zealand/20VR2549/2020\|EPI_ISL_579292\|2 | ···················· | ···················· | ···················· | ···················· | ···················· | ···················· |
| L | 2127 | hCoV-19/New Zealand/20VR2552/2020\|EPI_ISL_579294\|2 | ···················· | ···················· | ···················· | ···················· | ···················· | ···················· |
| L | 2128 | hCoV-19/New Zealand/20VR2555/2020\|EPI_ISL_579295\|2 | ···················· | ···················· | ···················· | ···················· | ···················· | ···················· |
| L | 2129 | hCoV-19/New Zealand/20VR2557/2020\|EPI_ISL_579296\|2 | ···················· | ···················· | ···················· | ···················· | ···················· | ···················· |
| L | 2130 | hCoV-19/New Zealand/20VR2558/2020\|EPI_ISL_579297\|2 | ···················· | ···················· | ···················· | ···················· | ···················· | ···················· |
| L | 2132 | hCoV-19/New Zealand/20VR2560/2020\|EPI_ISL_579299\|2 | ···················· | ···················· | ···················· | ···················· | ···················· | ···················· |
| L | 2133 | hCoV-19/England/ALDP-9EAA7E/2020\|EPI_ISL_580201\|20 | ···················· | ···················· | ···················· | ···················· | ···················· | ···················· |
| L | 2134 | hCoV-19/England/ALDP-9E9BC7/2020\|EPI_ISL_580202\|20 | ···················· | ···················· | ···················· | ···················· | ·····K·············· | ···················· |
| L | 2135 | hCoV-19/England/ALDP-9EA21C/2020\|EPI_ISL_580203\|20 | ···················· | ···················· | ···················· | ···················· | ···················· | ···················· |
| L | 2136 | hCoV-19/England/ALDP-9EAA9C/2020\|EPI_ISL_580204\|20 | ···················· | ···················· | ···················· | ···················· | ···················· | ···················· |
| L | 2137 | hCoV-19/England/ALDP-9E9B5E/2020\|EPI_ISL_580206\|20 | ···················· | ···················· | ···················· | ···················· | ···················· | ···················· |
| L | 2138 | hCoV-19/England/ALDP-9EA1F1/2020\|EPI_ISL_580207\|20 | ···················· | ·A·················· | ···················· | ···················· | ···················· | ···················· |
| L | 2139 | hCoV-19/England/ALDP-9EA1D3/2020\|EPI_ISL_580208\|20 | ···················· | ···················· | ···················· | ···················· | ···················· | ···················· |
| L | 2140 | hCoV-19/England/ALDP-9EAE72/2020\|EPI_ISL_580209\|20 | ···················· | ···················· | ···················· | ···················· | ···················· | ···················· |
| L | 2141 | hCoV-19/England/ALDP-9E9F16/2020\|EPI_ISL_580212\|20 | ···················· | ···················· | ···················· | ···················· | ···················· | ···················· |
| L | 2142 | hCoV-19/England/ALDP-9EA0A9/2020\|EPI_ISL_580214\|20 | ···················· | ···················· | ···················· | ···················· | ···················· | ···················· |
| L | 2143 | hCoV-19/England/ALDP-9EB07B/2020\|EPI_ISL_580215\|20 | ···················· | ···················· | ···················· | ···················· | ···················· | ···················· |
| L | 2144 | hCoV-19/England/ALDP-9EB06C/2020\|EPI_ISL_580216\|20 | ···················· | ···················· | ···················· | ···················· | ···················· | ···················· |
| L | 2145 | hCoV-19/England/ALDP-9EA4AD/2020\|EPI_ISL_580217\|20 | ···················· | ···················· | ···················· | ···················· | ···················· | ···················· |
| L | 2146 | hCoV-19/England/ALDP-9EA2C1/2020\|EPI_ISL_580218\|20 | ···················· | ···················· | ···················· | ···················· | ···················· | ···················· |
| L | 2147 | hCoV-19/England/ALDP-9EAC87/2020\|EPI_ISL_580219\|20 | ···················· | ···················· | ···················· | ···················· | ···················· | ···················· |
| L | 2148 | hCoV-19/England/ALDP-9E9F9E/2020\|EPI_ISL_580220\|20 | ···················· | ···················· | ···················· | ···················· | ···················· | ···················· |
| L | 2149 | hCoV-19/England/ALDP-9EB03F/2020\|EPI_ISL_580222\|20 | ···················· | ···················· | ···················· | ···················· | ···················· | ···················· |
| L | 2150 | hCoV-19/England/ALDP-9EADFD/2020\|EPI_ISL_580223\|20 | ···················· | ···················· | ···················· | ···················· | ···················· | ···················· |
| L | 2151 | hCoV-19/England/ALDP-9E79CF/2020\|EPI_ISL_580224\|20 | ···················· | ···················· | ···················· | ···················· | ···················· | ···················· |
| L | 2152 | hCoV-19/England/ALDP-9E9FE9/2020\|EPI_ISL_580226\|20 | ···················· | ···················· | ···················· | ···················· | ···················· | ···················· |
| L | 2153 | hCoV-19/England/ALDP-9EA07C/2020\|EPI_ISL_580227\|20 | ···················· | ···················· | ···················· | ···················· | ···················· | ···················· |
| L | 2154 | hCoV-19/England/ALDP-9EB099/2020\|EPI_ISL_580228\|20 | ···················· | ···················· | ···················· | ···················· | ···················· | ···················· |
| L | 2155 | hCoV-19/England/ALDP-9EA13D/2020\|EPI_ISL_580229\|20 | ···················· | ···················· | ···················· | ···················· | ···················· | ···················· |

L 2156 hCoV-19/England/ALDP-9E79ED/2020|EPI_ISL_580230|20 ···················· ·A·················· ···················· ···················· ···················· ····················
L 2157 hCoV-19/England/ALDP-9EAB11/2020|EPI_ISL_580232|20 ···················· ···················· ···················· ···················· ···················· ····················
L 2158 hCoV-19/England/ALDP-9EAE63/2020|EPI_ISL_580233|20 ···················· ···················· ···················· ···················· ···················· ····················
L 2159 hCoV-19/England/ALDP-9EA63E/2020|EPI_ISL_580235|20 ···················· ···················· ···················· ···················· ···················· ····················
L 2160 hCoV-19/England/ALDP-9EA188/2020|EPI_ISL_580236|20 ···················· ···················· ···················· ···················· ···················· ····················
L 2161 hCoV-19/England/ALDP-9EACD2/2020|EPI_ISL_580238|20 ···················· ···················· ···················· ···················· ···················· ····················
L 2162 hCoV-19/England/ALDP-9EA8DE/2020|EPI_ISL_580239|20 ···················· ···················· ···················· ···················· ···················· ····················
L 2163 hCoV-19/England/ALDP-9E76D7/2020|EPI_ISL_580240|20 ···················· ···················· ···················· ···················· ···················· ····················
L 2164 hCoV-19/England/ALDP-9EA838/2020|EPI_ISL_580241|20 ···················· ···················· ···················· ···················· ···················· ····················
L 2165 hCoV-19/England/ALDP-9EAAD8/2020|EPI_ISL_580242|20 ···················· ···················· ···················· ···················· ···················· ····················
L 2166 hCoV-19/England/ALDP-9E9F34/2020|EPI_ISL_580243|20 ···················· ···················· ···················· ···················· ···················· ····················
L 2167 hCoV-19/England/ALDP-9EA8CF/2020|EPI_ISL_580244|20 ···················· ···················· ···················· ···················· ···················· ····················
L 2168 hCoV-19/England/ALDP-9EA8A1/2020|EPI_ISL_580245|20 ···················· ···················· ···················· ···················· ···················· ····················
L 2169 hCoV-19/England/ALDP-9EADDF/2020|EPI_ISL_580246|20 ···················· ···················· ···················· ···················· ···················· ····················
L 2170 hCoV-19/England/ALDP-9EACC3/2020|EPI_ISL_580247|20 ···················· ···················· ···················· ···················· ···················· ····················
L 2171 hCoV-19/England/ALDP-9EB0A8/2020|EPI_ISL_580249|20 ···················· ···················· ···················· ···················· ···················· ····················
L 2172 hCoV-19/England/ALDP-9EAD66/2020|EPI_ISL_580251|20 ···················· ···················· ···················· ···················· ···················· ····················
L 2173 hCoV-19/England/ALDP-9E9AAC/2020|EPI_ISL_580252|20 ···················· ···················· ···················· ···················· ···················· ····················
L 2174 hCoV-19/England/ALDP-9EA49E/2020|EPI_ISL_580253|20 ···················· ···················· ···················· ···················· ···················· ····················
L 2175 hCoV-19/England/ALDP-9E9A9D/2020|EPI_ISL_580254|20 ···················· ···················· ···················· ···················· ···················· ····················
L 2176 hCoV-19/England/ALDP-9E79FC/2020|EPI_ISL_580255|20 ···················· ···················· ···················· ···················· ···················· ····················
L 2177 hCoV-19/England/ALDP-9E9B6D/2020|EPI_ISL_580257|20 ···················· ·A·················· ···················· ···················· ···················· ··A·················
L 2178 hCoV-19/England/ALDP-9EA4BC/2020|EPI_ISL_580258|20 ···················· ···················· ···················· ···················· ···················· ····················
L 2179 hCoV-19/Wales/ALDP-9EABC6/2020|EPI_ISL_580260|2020 ···················· ···················· ···················· ···················· ···················· ····················
L 2180 hCoV-19/England/ALDP-9EAC69/2020|EPI_ISL_580261|20 ···················· ···················· ···················· ···················· ···················· ····················
L 2181 hCoV-19/England/ALDP-9E9A60/2020|EPI_ISL_580262|20 ···················· ···················· ···················· ···················· ···················· ····················
L 2182 hCoV-19/England/ALDP-9EA11F/2020|EPI_ISL_580263|20 ···················· ···················· ···················· ···················· ···················· ····················
L 2183 hCoV-19/England/ALDP-9E79A1/2020|EPI_ISL_580264|20 ···················· ···················· ···················· ···················· ···················· ····················
L 2184 hCoV-19/England/ALDP-9EA962/2020|EPI_ISL_580265|20 ···················· ···················· ···················· ···················· ···················· ····················
L 2185 hCoV-19/England/ALDP-9EAEAF/2020|EPI_ISL_580266|20 ···················· ···················· ···················· ···················· ···················· ····················
L 2186 hCoV-19/England/ALDP-9E9E46/2020|EPI_ISL_580267|20 ···················· ···················· ···················· ···················· ···················· ····················
L 2187 hCoV-19/England/ALDP-9E9A8E/2020|EPI_ISL_580268|20 ···················· ···················· ···················· ···················· ···················· ····················
L 2188 hCoV-19/England/ALDP-9EAF51/2020|EPI_ISL_580269|20 ···················· ···················· ···················· ···················· ···················· ····················
L 2189 hCoV-19/England/ALDP-9E9B4F/2020|EPI_ISL_580270|20 ···················· ·A·················· ···················· ···················· ···················· ····················
L 2190 hCoV-19/England/ALDP-9EA3EC/2020|EPI_ISL_580271|20 ···················· ···················· ···················· ···················· ···················· ····················
L 2191 hCoV-19/England/ALDP-9EA601/2020|EPI_ISL_580273|20 ···················· ···················· ···················· ···················· ···················· ····················
L 2192 hCoV-19/England/ALDP-9EAAF6/2020|EPI_ISL_580274|20 ···················· ···················· ···················· ···················· ···················· ····················
L 2193 hCoV-19/England/ALDP-9E62D4/2020|EPI_ISL_580275|20 ···················· ···················· ···················· ···················· ···················· ····················
L 2194 hCoV-19/England/ALDP-9E6359/2020|EPI_ISL_580276|20 ···················· ···················· ···················· ···················· ···················· ····················
L 2195 hCoV-19/England/ALDP-9E69FD/2020|EPI_ISL_580277|20 ···················· ···················· ···················· ···················· ···················· ····················
L 2196 hCoV-19/England/ALDP-9E6799/2020|EPI_ISL_580278|20 ···················· ·A·················· ···················· ···················· ···················· ··A·················
L 2197 hCoV-19/England/ALDP-9E719A/2020|EPI_ISL_580279|20 ···················· ···················· ···················· ···················· ···················· ····················
L 2198 hCoV-19/England/ALDP-9E634A/2020|EPI_ISL_580280|20 ···················· ···················· ···················· ···················· ···················· ····················
L 2199 hCoV-19/England/CAMC-9EC502/2020|EPI_ISL_580282|20 ···················· ···················· ···················· ···················· ···················· ····················
L 2200 hCoV-19/England/CAMC-9EC739/2020|EPI_ISL_580283|20 ···················· ·A·······A·········· ···················· ···················· ···················· ····················
L 2201 hCoV-19/England/ALDP-9E6BE8/2020|EPI_ISL_580284|20 ···················· ···················· ···················· ···················· ···················· ····················
L 2202 hCoV-19/USA/VA-DCLS-1534/2020|EPI_ISL_581530|2020- ···················· ···················· ···················· ···················· ···················· ····················
L 2203 hCoV-19/USA/VA-DCLS-1536/2020|EPI_ISL_581531|2020- ···················· ·A·················· ···················· ···················· ···················· ····················
L 2204 hCoV-19/USA/VA-DCLS-1537/2020|EPI_ISL_581532|2020- ···················· ···················· ···················· ···················· ···················· ····················
L 2205 hCoV-19/USA/VA-DCLS-1539/2020|EPI_ISL_581534|2020- ···················· ···················· ···················· ···················· ···················· ····················
L 2206 hCoV-19/USA/VA-DCLS-1540/2020|EPI_ISL_581535|2020- ···················· ···················· ···················· ···················· ···················· ····················
L 2207 hCoV-19/USA/VA-DCLS-1545/2020|EPI_ISL_581536|2020- ···················· ···················· ···················· ···················· ···················· ····················
L 2208 hCoV-19/USA/VA-DCLS-1546/2020|EPI_ISL_581537|2020- ···················· ···················· ···················· ···················· ···················· ····················
L 2209 hCoV-19/USA/VA-DCLS-1547/2020|EPI_ISL_581538|2020- ···················· ·A·················· ···················· ···················· ···················· ····················
L 2210 hCoV-19/USA/VA-DCLS-1548/2020|EPI_ISL_581539|2020- ···················· ·A·················· ···················· ···················· ···················· ····················
L 2211 hCoV-19/USA/VA-DCLS-1550/2020|EPI_ISL_581540|2020- ···················· ·A·················· ···················· ···················· ···················· ····················
L 2212 hCoV-19/USA/VA-DCLS-1552/2020|EPI_ISL_581541|2020- ···················· ···················· ···················· ···················· ···················· ····················
L 2213 hCoV-19/USA/VA-DCLS-1416/2020|EPI_ISL_581542|2020- ···················· ···················· ···················· ···················· ···················· ····················
L 2214 hCoV-19/USA/VA-DCLS-1420/2020|EPI_ISL_581543|2020- ···················· ···················· ···················· ···················· ···················· ····················
L 2215 hCoV-19/USA/VA-DCLS-1421/2020|EPI_ISL_581544|2020- ···················· ···················· ···················· ···················· ···················· ····················
L 2216 hCoV-19/USA/VA-DCLS-1422/2020|EPI_ISL_581545|2020- ···················· ···················· ···················· ···················· ···················· ·········A··········
L 2217 hCoV-19/USA/VA-DCLS-1423/2020|EPI_ISL_581546|2020- ···················· ···················· ···················· ···················· ···················· ····················
L 2218 hCoV-19/USA/VA-DCLS-1424/2020|EPI_ISL_581547|2020- ···················· ···················· ···················· ···················· ···················· ····················
L 2219 hCoV-19/USA/VA-DCLS-1425/2020|EPI_ISL_581548|2020- ···················· ···················· ···················· ···················· ···················· ····················
L 2220 hCoV-19/USA/VA-DCLS-1426/2020|EPI_ISL_581549|2020- ···················· ···················· ···················· ···················· ···················· ····················
L 2221 hCoV-19/USA/VA-DCLS-1427/2020|EPI_ISL_581550|2020- ···················· ···················· ···················· ···················· ···················· ····················
L 2222 hCoV-19/USA/VA-DCLS-1429/2020|EPI_ISL_581551|2020- ···················· ···················· ···················· ···················· ···················· ····················
L 2223 hCoV-19/USA/VA-DCLS-1430/2020|EPI_ISL_581552|2020- ···················· ···················· ···················· ···················· ···················· ····················
L 2224 hCoV-19/USA/VA-DCLS-1431/2020|EPI_ISL_581553|2020- ···················· ···················· ···················· ···················· ···················· ····················
L 2225 hCoV-19/USA/VA-DCLS-1432/2020|EPI_ISL_581554|2020- ···················· ···················· ···················· ···················· ···················· ····················
L 2226 hCoV-19/USA/VA-DCLS-1433/2020|EPI_ISL_581555|2020- ···················· ···················· ···················· ···················· ···················· ····················
L 2227 hCoV-19/USA/VA-DCLS-1434/2020|EPI_ISL_581556|2020- ···················· ···················· ···················· ···················· ···················· ····················
L 2228 hCoV-19/USA/VA-DCLS-1436/2020|EPI_ISL_581557|2020- ···················· ···················· ···················· ···················· ···················· ····················
L 2229 hCoV-19/USA/VA-DCLS-1481/2020|EPI_ISL_581558|2020- ···················· ···················· ···················· ···················· ···················· ····················
L 2230 hCoV-19/USA/VA-DCLS-1482/2020|EPI_ISL_581559|2020- ···················· ···················· ···················· ···················· ···················· ····················
L 2231 hCoV-19/USA/VA-DCLS-1484/2020|EPI_ISL_581561|2020- ···················· ···················· ···················· ···················· ···················· ····················
L 2232 hCoV-19/USA/VA-DCLS-1485/2020|EPI_ISL_581562|2020- ···················· ···················· ···················· ···················· ···················· ····················
L 2233 hCoV-19/USA/VA-DCLS-1487/2020|EPI_ISL_581563|2020- ···················· ···················· ···················· ···················· ···················· ····················
L 2234 hCoV-19/USA/VA-DCLS-1490/2020|EPI_ISL_581564|2020- ···················· ···················· ···················· ···················· ···················· ····················
L 2235 hCoV-19/USA/VA-DCLS-1491/2020|EPI_ISL_581565|2020- ···················· ···················· ···················· ···················· ···················· ····················

L 2236 hCoV-19/USA/VA-DCLS-1492/2020|EPI_ISL_581566|2020- ···················· ·A·················· ···················· ···················· ···················· ····················
L 2237 hCoV-19/USA/VA-DCLS-1493/2020|EPI_ISL_581567|2020- ···················· ···················· ···················· ···················· ···················· ····················
L 2238 hCoV-19/USA/VA-DCLS-1495/2020|EPI_ISL_581568|2020- ···················· ···················· ···················· ···················· ···················· ····················
L 2239 hCoV-19/USA/VA-DCLS-1497/2020|EPI_ISL_581569|2020- ···················· ···················· ···················· ···················· ···················· ····················
L 2240 hCoV-19/USA/VA-DCLS-1499/2020|EPI_ISL_581570|2020- ···················· ···················· ···················· ···················· ···················· ····················
L 2241 hCoV-19/USA/VA-DCLS-1502/2020|EPI_ISL_581571|2020- ···················· ···A················ ···················· ···················· ···················· ····················
L 2242 hCoV-19/USA/VA-DCLS-1504/2020|EPI_ISL_581573|2020- ···················· ···················· ···················· ···················· ···················· ····················
L 2243 hCoV-19/USA/VA-DCLS-1505/2020|EPI_ISL_581574|2020- ···················· ···················· ···················· ···················· ···················· ····················
L 2244 hCoV-19/Belgium/ULG-10428/2020|EPI_ISL_581586|2020 ···················· ···················· ···················· ···················· ···················· ····················
L 2249 hCoV-19/Belgium/ULG-10413/2020|EPI_ISL_581595|2020 ···················· ···················· ···················· ···················· ···················· ····················
L 2250 hCoV-19/Belgium/ULG-10414/2020|EPI_ISL_581596|2020 ···················· ···················· ···················· ···················· ···················· ····················
L 2251 hCoV-19/Belgium/ULG-10415/2020|EPI_ISL_581597|2020 ···················· ···················· ···················· ···················· ···················· ····················
L 2252 hCoV-19/Belgium/ULG-10416/2020|EPI_ISL_581598|2020 ···················· ···················· ···················· ···················· ···················· ····················
L 2253 hCoV-19/Belgium/ULG-10417/2020|EPI_ISL_581599|2020 ···················· ···················· ···················· ···················· ···················· ····················
L 2254 hCoV-19/Sweden/20-52366/2020|EPI_ISL_582800|2020-0 ···················· ···················· ···················· ···················· ···················· ····················
L 2255 hCoV-19/Sweden/20-52534/2020|EPI_ISL_582801|2020-0 ···················· ···················· ···················· ···················· ···················· ····················
L 2256 hCoV-19/Sweden/20-52671/2020|EPI_ISL_582803|2020-0 ···················· ···················· ···················· ···················· ···················· ····················
L 2257 hCoV-19/Sweden/20-52672/2020|EPI_ISL_582804|2020-0 ···················· ···················· ···················· ···················· ···················· ····················
L 2258 hCoV-19/Sweden/20-52673/2020|EPI_ISL_582805|2020-0 ···················· ···················· ···················· ···················· ···················· ····················
L 2259 hCoV-19/Sweden/20-52684/2020|EPI_ISL_582806|2020-0 ···················· ···················· ···················· ···················· ···················· ····················
L 2260 hCoV-19/Sweden/20-52686/2020|EPI_ISL_582807|2020-0 ···················· ···················· ···················· ···················· ···················· ····················
L 2261 hCoV-19/Sweden/20-52783/2020|EPI_ISL_582808|2020-0 ···················· ···················· ···················· ···················· ···················· ····················
L 2262 hCoV-19/Sweden/20-52800/2020|EPI_ISL_582809|2020-0 ···················· ···················· ···················· ···················· ···················· ····················
L 2263 hCoV-19/Italy/SIC-IZSBM169-59/2020|EPI_ISL_582810| ···················· ···················· ···················· ···················· ···················· ····················
L 2264 hCoV-19/Italy/SIC-IZSBM169-60/2020|EPI_ISL_582811| ···················· ···················· ···················· ···················· ···················· ····················
L 2265 hCoV-19/USA/CA-CZB-2754/2020|EPI_ISL_582860|2020-0 ···················· ·A·················· ···················· ···················· ···················· ····················
L 2266 hCoV-19/USA/CA-CZB-2755/2020|EPI_ISL_582861|2020-0 ···················· ·A·················· ···················· ···················· ···················· ····················
L 2267 hCoV-19/USA/CA-CZB-2757/2020|EPI_ISL_582862|2020-0 ···················· ·A·················· ···················· ···················· ···················· ····················
L 2268 hCoV-19/USA/CA-CZB-2758/2020|EPI_ISL_582863|2020-0 ···················· ·A·················· ···················· ···················· ···················· ····················
L 2269 hCoV-19/USA/CA-CZB-2759/2020|EPI_ISL_582864|2020-0 ···················· ·A·················· ···················· ···················· ···················· ····················
L 2270 hCoV-19/USA/CA-CZB-2760/2020|EPI_ISL_582865|2020-0 ···················· ·A·················· ···················· ···················· ···················· ····················
L 2271 hCoV-19/USA/CA-CZB-2761/2020|EPI_ISL_582866|2020-0 ···················· ·A·················· ···················· ···················· ···················· ····················
L 2272 hCoV-19/USA/CA-CZB-2763/2020|EPI_ISL_582867|2020-0 ···················· ·A·················· ···················· ···················· ···················· ····················
L 2273 hCoV-19/USA/CA-CZB-2764/2020|EPI_ISL_582868|2020-0 ···················· ·A·················· ···················· ···················· ···················· ····················
L 2274 hCoV-19/USA/CA-CZB-2770/2020|EPI_ISL_582869|2020-0 ···················· ·A·················· ···················· ···················· ···················· ····················
L 2275 hCoV-19/USA/CA-CZB-2771/2020|EPI_ISL_582870|2020-0 ···················· ·A·················· ···················· ···················· ···················· ····················
L 2276 hCoV-19/USA/CA-CZB-2794/2020|EPI_ISL_582871|2020-0 ···················· ·A·················· ···················· ···················· ···················· ····················
L 2277 hCoV-19/USA/CA-CZB-2806/2020|EPI_ISL_582872|2020-0 ···················· ·A·················· ···················· ···················· ···················· ····················
L 2278 hCoV-19/USA/CA-CZB-2807/2020|EPI_ISL_582873|2020-0 ···················· ·A·················· ···················· ···················· ···················· ····················
L 2279 hCoV-19/USA/CA-CZB-2808/2020|EPI_ISL_582874|2020-0 ···················· ·A·················· ···················· ···················· ···················· ····················
L 2280 hCoV-19/USA/CA-CZB-2810/2020|EPI_ISL_582875|2020-0 ···················· ·A·················· ···················· ···················· ···················· ····················
L 2281 hCoV-19/USA/CA-CZB-2813/2020|EPI_ISL_582876|2020-0 ···················· ·A·················· ···················· ···················· ···················· ····················
L 2282 hCoV-19/USA/CA-CZB-2814/2020|EPI_ISL_582877|2020-0 ···················· ·A·················· ···················· ···················· ···················· ····················
L 2283 hCoV-19/USA/CA-CZB-2815/2020|EPI_ISL_582878|2020-0 ···················· ·A·················· ···················· ···················· ···················· ····················
L 2284 hCoV-19/USA/CA-CZB-2829/2020|EPI_ISL_582879|2020-0 ···················· ·A·················· ···················· ···················· ···················· ····················
L 2285 hCoV-19/USA/CA-CZB-6802/2020|EPI_ISL_582880|2020-0 ···················· ·A·················· ···················· ···················· ···················· ····················
L 2286 hCoV-19/USA/CA-CZB-6804/2020|EPI_ISL_582881|2020-0 ···················· ·A·················· ···················· ···················· ···················· ····················
L 2287 hCoV-19/USA/CA-CZB-6805/2020|EPI_ISL_582882|2020-0 ···················· ·A·················· ···················· ···················· ···················· ····················
L 2288 hCoV-19/USA/CA-CZB-6807/2020|EPI_ISL_582883|2020-0 ···················· ·A·················· ···················· ···················· ···················· ····················
L 2289 hCoV-19/USA/CA-CZB-6809/2020|EPI_ISL_582884|2020-0 ···················· ·A·················· ···················· ···················· ···················· ····················
L 2290 hCoV-19/USA/CA-CZB-6810/2020|EPI_ISL_582885|2020-0 ···················· ·A·················· ···················· ···················· ···················· ····················
L 2291 hCoV-19/USA/CA-CZB-6811/2020|EPI_ISL_582886|2020-0 ···················· ·A·················· ···················· ···················· ···················· ····················
L 2292 hCoV-19/USA/CA-CZB-6927/2020|EPI_ISL_582887|2020-0 ···················· ···················· ···················· ···················· ···················· ····················
L 2293 hCoV-19/USA/CA-CZB-6928/2020|EPI_ISL_582888|2020-0 ···················· ···················· ···················· ···················· ···················· ····················
L 2294 hCoV-19/USA/CA-CZB-6929/2020|EPI_ISL_582889|2020-0 ···················· ···················· ···················· ···················· ···················· ····················
L 2295 hCoV-19/USA/CA-CZB-6932/2020|EPI_ISL_582890|2020-0 ···················· ···················· ···················· ···················· ···················· ····················
L 2296 hCoV-19/USA/CA-CZB-6933/2020|EPI_ISL_582891|2020-0 ···················· ·A·················· ···················· ···················· ···················· ····················
L 2297 hCoV-19/USA/CA-CZB-6938/2020|EPI_ISL_582892|2020-0 ···················· ·A·················· ···················· ···················· ···················· ····················
L 2298 hCoV-19/USA/CA-CZB-6941/2020|EPI_ISL_582893|2020-0 ···················· ·A·················· ···················· ···················· ···················· ····················
L 2299 hCoV-19/USA/CA-CZB-4714/2020|EPI_ISL_582894|2020-0 ···················· ···················· ···················· ···················· ···················· ····················
L 2300 hCoV-19/USA/CA-CZB-4730/2020|EPI_ISL_582895|2020-0 ···················· ·A·················· ···················· ···················· ···················· ····················
L 2301 hCoV-19/USA/CA-CZB-4740/2020|EPI_ISL_582896|2020-0 ···················· ·A·················· ···················· ···················· ···················· ····················
L 2302 hCoV-19/USA/CA-CZB-4812/2020|EPI_ISL_582897|2020-0 ···················· ·A·················· ···················· ···················· ···················· ····················
L 2303 hCoV-19/USA/CA-CZB-6964/2020|EPI_ISL_582899|2020-0 ···················· ·A·················· ···················· ···················· ···················· ····················
L 2304 hCoV-19/Australia/VIC7904/2020|EPI_ISL_594134|2020 ···················· ···················· ···················· ···················· ···················· ····················
L 2305 hCoV-19/England/ALDP-9BCE5B/2020|EPI_ISL_573080|20 ···················· ···················· ···················· ···················· ···················· ····················
L 2306 hCoV-19/England/ALDP-9BCF2B/2020|EPI_ISL_573081|20 ···················· ···················· ···················· ···················· ···················· ····················
L 2307 hCoV-19/England/ALDP-9BCF49/2020|EPI_ISL_573082|20 ···················· ···················· ···················· ···················· ···················· ····················
L 2308 hCoV-19/England/ALDP-9CC16E/2020|EPI_ISL_573088|20 ···················· ···················· ···················· ···················· ···················· ····················
L 2312 hCoV-19/England/ALDP-9CC5CC/2020|EPI_ISL_573116|20 ···················· ···················· ···················· ···················· ···················· ····················
L 2313 hCoV-19/England/ALDP-9CC5EA/2020|EPI_ISL_573117|20 ···················· ···················· ···················· ···················· ···················· ····················
L 2314 hCoV-19/England/ALDP-9CC5F9/2020|EPI_ISL_573118|20 ···················· ···················· ···················· ···················· ···················· ····················
L 2315 hCoV-19/England/ALDP-9CC632/2020|EPI_ISL_573119|20 ···················· ·A·················· ···················· ···················· ···················· ··A·················
L 2316 hCoV-19/England/ALDP-9CC66F/2020|EPI_ISL_573120|20 ···················· ·A·················· ···················· ···················· ···················· ··A·················
L 2318 hCoV-19/England/ALDP-9CC975/2020|EPI_ISL_573122|20 ···················· ···················· ···················· ···················· ···················· ····················
L 2319 hCoV-19/England/ALDP-9CC9EE/2020|EPI_ISL_573123|20 ···················· ···················· ···················· ···················· ···················· ····················
L 2322 hCoV-19/England/ALDP-9CCADC/2020|EPI_ISL_573126|20 ···················· ···················· ···················· ···················· ···················· ····················
L 2323 hCoV-19/England/ALDP-9CCAEB/2020|EPI_ISL_573127|20 ···················· ···················· ···················· ···················· ···················· ····················
L 2328 hCoV-19/England/CAMB-1B5E4B/2020|EPI_ISL_573132|20 ···················· ···················· ···················· ···················· ···················· ····················
L 2329 hCoV-19/England/CAMB-1B5E69/2020|EPI_ISL_573133|20 ···················· ···················· ···················· ···················· ···················· ····················

| | | | | | | | |
|---|---|---|---|---|---|---|---|
| L | 2330 hCoV-19/England/CAMB-1B5E78/2020\|EPI_ISL_573134\|20 | ···················· | ···················· | ···················· | ···················· | ···················· | ···················· |
| L | 2331 hCoV-19/England/CAMB-1B5E87/2020\|EPI_ISL_573135\|20 | ···················· | ···················· | ···················· | ···················· | ···················· | ···················· |
| L | 2332 hCoV-19/England/CAMB-1B5EC3/2020\|EPI_ISL_573136\|20 | ···················· | ···················· | ···················· | ···················· | ···················· | ···················· |
| L | 2333 hCoV-19/England/CAMB-1B5ED2/2020\|EPI_ISL_573137\|20 | ···················· | ···················· | ···················· | ···················· | ···················· | ···················· |
| L | 2334 hCoV-19/England/CAMB-1B5F2A/2020\|EPI_ISL_573138\|20 | ···················· | ···················· | ···················· | ···················· | ···················· | ···················· |
| L | 2335 hCoV-19/England/CAMB-1B5F39/2020\|EPI_ISL_573139\|20 | ···················· | ···················· | ···················· | ···················· | ···················· | ···················· |
| L | 2336 hCoV-19/England/CAMB-1B5F48/2020\|EPI_ISL_573140\|20 | ···················· | ···················· | ···················· | ···················· | ···················· | ···················· |
| L | 2339 hCoV-19/England/CAMB-1B5F84/2020\|EPI_ISL_573143\|20 | ···················· | ···················· | ···················· | ···················· | ···················· | ···················· |
| L | 2340 hCoV-19/England/CAMB-1B6035/2020\|EPI_ISL_573144\|20 | ···················· | ···················· | ···················· | ···················· | ···················· | ···················· |
| L | 2341 hCoV-19/England/CAMB-1B6114/2020\|EPI_ISL_573145\|20 | ···················· | ···················· | ···················· | ···················· | ···················· | ···················· |
| L | 2342 hCoV-19/England/CAMB-1B6132/2020\|EPI_ISL_573146\|20 | ···················· | ···················· | ···················· | ···················· | ···················· | ···················· |
| L | 2346 hCoV-19/England/CAMB-1B617E/2020\|EPI_ISL_573150\|20 | ···················· | ···················· | ···················· | ···················· | ···················· | ···················· |
| L | 2347 hCoV-19/England/CAMB-1B618D/2020\|EPI_ISL_573151\|20 | ···················· | ···················· | ···················· | ···················· | ···················· | ···················· |
| L | 2348 hCoV-19/Germany/NW-HHU-D871/2020\|EPI_ISL_573152\|20 | ···················· | ···················· | ···················· | ···················· | ···················· | ···················· |
| L | 2350 hCoV-19/England/CAMC-9BFE2B/2020\|EPI_ISL_573154\|20 | ···················· | ···················· | ···················· | ···················· | ···················· | ···················· |
| L | 2351 hCoV-19/England/CAMC-9BFE49/2020\|EPI_ISL_573155\|20 | ···················· | ···················· | ···················· | ···················· | ···················· | ···················· |
| L | 2352 hCoV-19/England/CAMC-9BFE58/2020\|EPI_ISL_573156\|20 | ···················· | ·A·················· | ···················· | ···················· | ···················· | ···················· |
| L | 2353 hCoV-19/England/CAMC-9BFE76/2020\|EPI_ISL_573157\|20 | ···················· | ···················· | ···················· | ···················· | ···················· | ···················· |
| L | 2354 hCoV-19/England/CAMC-9BFE85/2020\|EPI_ISL_573158\|20 | ···················· | ·A·······A·········· | ···················· | ···················· | ···················· | ···················· |
| L | 2355 hCoV-19/England/CAMC-9BFEC1/2020\|EPI_ISL_573159\|20 | ···················· | ···················· | ···················· | ···················· | ···················· | ···················· |
| L | 2356 hCoV-19/England/CAMC-9BFEEF/2020\|EPI_ISL_573160\|20 | ···················· | ···················· | ···················· | ···················· | ···················· | ···················· |
| L | 2357 hCoV-19/England/CAMC-9BFF0A/2020\|EPI_ISL_573161\|20 | ···················· | ···················· | ···················· | ···················· | ···················· | ···················· |
| L | 2358 hCoV-19/England/CAMC-9BFFA0/2020\|EPI_ISL_573162\|20 | ···················· | ···················· | ···················· | ···················· | ···················· | ···················· |
| L | 2359 hCoV-19/England/CAMC-9BFFBF/2020\|EPI_ISL_573163\|20 | ···················· | ···················· | ···················· | ···················· | ···················· | ···················· |
| L | 2360 hCoV-19/England/CAMC-9BFFEC/2020\|EPI_ISL_573164\|20 | ···················· | ···················· | ···················· | ···················· | ···················· | ···················· |
| L | 2361 hCoV-19/England/CAMC-9BFFFB/2020\|EPI_ISL_573165\|20 | ···················· | ···················· | ···················· | ···················· | ···················· | ···················· |
| L | 2362 hCoV-19/England/CAMC-9C0003/2020\|EPI_ISL_573166\|20 | ···················· | ···················· | ···················· | ···················· | ···················· | ···················· |
| L | 2363 hCoV-19/England/CAMC-9C005E/2020\|EPI_ISL_573167\|20 | ···················· | ·A·················· | ···················· | ···················· | ···················· | ···A················ |
| L | 2364 hCoV-19/England/CAMC-9C006D/2020\|EPI_ISL_573168\|20 | ···················· | ···················· | ···················· | ···················· | ···················· | ···················· |
| L | 2365 hCoV-19/England/CAMC-9C00D6/2020\|EPI_ISL_573169\|20 | ···················· | ···················· | ···················· | ···················· | ···················· | ···················· |
| L | 2366 hCoV-19/England/CAMC-9C011F/2020\|EPI_ISL_573170\|20 | ···················· | ···················· | ···················· | ···················· | ······T············· | ···················· |
| L | 2367 hCoV-19/England/CAMC-9C014C/2020\|EPI_ISL_573171\|20 | ···················· | ···················· | ···················· | ···················· | ···················· | ···················· |
| L | 2368 hCoV-19/England/CAMC-9C015B/2020\|EPI_ISL_573172\|20 | ···················· | ···················· | ···················· | ···················· | ···················· | ···················· |
| L | 2369 hCoV-19/England/CAMC-9C01A6/2020\|EPI_ISL_573173\|20 | ···················· | ···················· | ···················· | ···················· | ···················· | ···················· |
| L | 2370 hCoV-19/England/CAMC-9C01B5/2020\|EPI_ISL_573174\|20 | ···················· | ···················· | ···················· | ···················· | ···················· | ···················· |
| L | 2371 hCoV-19/England/CAMC-9C01E2/2020\|EPI_ISL_573175\|20 | ···················· | ···················· | ···················· | ···················· | ···················· | ···················· |
| L | 2372 hCoV-19/England/CAMC-9C0258/2020\|EPI_ISL_573176\|20 | ···················· | ···················· | ···················· | ···················· | ···················· | ···················· |
| L | 2373 hCoV-19/England/CAMC-9C0294/2020\|EPI_ISL_573177\|20 | ···················· | ···················· | ···················· | ···················· | ···················· | ···················· |
| L | 2374 hCoV-19/England/CAMC-9C02C1/2020\|EPI_ISL_573178\|20 | ···················· | ·A·················· | ···················· | ···················· | ···················· | ···················· |
| L | 2375 hCoV-19/England/CAMC-9C0319/2020\|EPI_ISL_573179\|20 | ···················· | ···················· | ···················· | ···················· | ···················· | ···················· |
| L | 2376 hCoV-19/England/CAMC-9C0364/2020\|EPI_ISL_573180\|20 | ···················· | ···················· | ···················· | ···················· | ···················· | ···················· |
| L | 2377 hCoV-19/England/CAMC-9C0373/2020\|EPI_ISL_573181\|20 | ···················· | ···················· | ···················· | ···················· | ···················· | ···················· |
| L | 2378 hCoV-19/England/CAMC-9C0382/2020\|EPI_ISL_573182\|20 | ···················· | ···················· | ···················· | ···················· | ···················· | ···················· |
| L | 2379 hCoV-19/England/CAMC-9C03A0/2020\|EPI_ISL_573183\|20 | ···················· | ······C············· | ···················· | ···················· | ···················· | ···················· |
| L | 2380 hCoV-19/England/CAMC-9C03DD/2020\|EPI_ISL_573184\|20 | ···················· | ···················· | ···················· | ···················· | ···················· | ···················· |
| L | 2381 hCoV-19/England/CAMC-9C0425/2020\|EPI_ISL_573185\|20 | ···················· | ···················· | ···················· | ···················· | ···················· | ···················· |
| L | 2382 hCoV-19/England/CAMC-9C0452/2020\|EPI_ISL_573186\|20 | ···················· | ···················· | ···················· | ···················· | ···················· | ···················· |
| L | 2383 hCoV-19/England/CAMC-9C048F/2020\|EPI_ISL_573187\|20 | ···················· | ···················· | ···················· | ···················· | ···················· | ···················· |
| L | 2384 hCoV-19/England/CAMC-9C0540/2020\|EPI_ISL_573188\|20 | ···················· | ···················· | ···················· | ···················· | ···················· | ···················· |
| L | 2385 hCoV-19/England/CAMC-9C056E/2020\|EPI_ISL_573189\|20 | ···················· | ···················· | ···················· | ···················· | ···················· | ···················· |
| L | 2386 hCoV-19/England/CAMC-9C057D/2020\|EPI_ISL_573190\|20 | ···················· | ···················· | ···················· | ···················· | ···················· | ···················· |
| L | 2387 hCoV-19/England/CAMC-9C05AA/2020\|EPI_ISL_573191\|20 | ···················· | ···················· | ···················· | ···················· | ···················· | ···················· |
| L | 2388 hCoV-19/England/CAMC-9C062F/2020\|EPI_ISL_573192\|20 | ···················· | ···················· | ···················· | ···················· | ···················· | ···················· |
| L | 2389 hCoV-19/England/CAMC-9C0768/2020\|EPI_ISL_573193\|20 | ···················· | ···················· | ···················· | ···················· | ···················· | ···················· |
| L | 2390 hCoV-19/England/CAMC-9C0962/2020\|EPI_ISL_573194\|20 | ···················· | ····A··············· | ···················· | ···················· | ···················· | ···················· |
| L | 2391 hCoV-19/England/CAMC-9C163D/2020\|EPI_ISL_573195\|20 | ···················· | ···················· | ···················· | ···················· | ···················· | ···················· |
| L | 2392 hCoV-19/England/CAMC-9C16A6/2020\|EPI_ISL_573196\|20 | ···················· | ·A·················· | ···················· | ···················· | ···················· | ···················· |
| L | 2393 hCoV-19/England/CAMC-9C16B5/2020\|EPI_ISL_573197\|20 | ···················· | ···················· | ···················· | ···················· | ···················· | ···················· |
| L | 2394 hCoV-19/England/CAMC-9C172B/2020\|EPI_ISL_573198\|20 | ···················· | ···················· | ···················· | ···················· | ···················· | ···················· |
| L | 2395 hCoV-19/England/CAMC-9C173A/2020\|EPI_ISL_573199\|20 | ···················· | ···················· | ···················· | ···················· | ···················· | ···················· |
| L | 2396 hCoV-19/USA/TX-DSHS-0541/2020\|EPI_ISL_574445\|2020- | ···················· | ···················· | ···················· | ···················· | ···················· | ···················· |
| L | 2397 hCoV-19/USA/TX-DSHS-0542/2020\|EPI_ISL_574446\|2020- | ···················· | ···················· | ···················· | ···················· | ···················· | ···················· |
| L | 2398 hCoV-19/USA/TX-DSHS-0543/2020\|EPI_ISL_574447\|2020- | ···················· | ···················· | ···················· | ···················· | ···················· | ···················· |
| L | 2399 hCoV-19/USA/TX-DSHS-0544/2020\|EPI_ISL_574448\|2020- | ···················· | ···················· | ···················· | ···················· | ···················· | ···················· |
| L | 2400 hCoV-19/USA/TX-DSHS-0548/2020\|EPI_ISL_574452\|2020- | ···················· | ···················· | ···················· | ···················· | ···················· | ···················· |
| L | 2401 hCoV-19/USA/TX-DSHS-0550/2020\|EPI_ISL_574454\|2020- | ···················· | ···················· | ···················· | ···················· | ···················· | ···················· |
| L | 2402 hCoV-19/USA/TX-DSHS-0553/2020\|EPI_ISL_574456\|2020- | ···················· | ···················· | ···················· | ···················· | ···················· | ···················· |
| L | 2403 hCoV-19/USA/TX-DSHS-0554/2020\|EPI_ISL_574457\|2020- | ···················· | ···················· | ···················· | ···················· | ···················· | ···················· |
| L | 2404 hCoV-19/USA/TX-DSHS-0555/2020\|EPI_ISL_574458\|2020- | ···················· | ···················· | ···················· | ···················· | ···················· | ···················· |
| L | 2405 hCoV-19/USA/TX-DSHS-0557/2020\|EPI_ISL_574460\|2020- | ···················· | ···················· | ···················· | ···················· | ···················· | ···················· |
| L | 2406 hCoV-19/USA/TX-DSHS-0558/2020\|EPI_ISL_574461\|2020- | ···················· | ···················· | ···················· | ···················· | ···················· | ···················· |
| L | 2407 hCoV-19/USA/TX-DSHS-0559/2020\|EPI_ISL_574462\|2020- | ···················· | ···················· | ···················· | ···················· | ···················· | ···················· |
| L | 2408 hCoV-19/USA/TX-DSHS-0560/2020\|EPI_ISL_574463\|2020- | ···················· | ···················· | ···················· | ···················· | ···················· | ···················· |
| L | 2409 hCoV-19/USA/TX-DSHS-0562/2020\|EPI_ISL_574464\|2020- | ···················· | ···················· | ···················· | ···················· | ···················· | ···················· |
| L | 2410 hCoV-19/USA/TX-DSHS-0563/2020\|EPI_ISL_574465\|2020- | ···················· | ···················· | ···················· | ···················· | ···················· | ···················· |
| L | 2411 hCoV-19/USA/TX-DSHS-0564/2020\|EPI_ISL_574466\|2020- | ···················· | ···················· | ···················· | ···················· | ···················· | ···················· |
| L | 2412 hCoV-19/USA/TX-DSHS-0565/2020\|EPI_ISL_574467\|2020- | ···················· | ···················· | ···················· | ···················· | ···················· | ···················· |
| L | 2413 hCoV-19/USA/TX-DSHS-0566/2020\|EPI_ISL_574468\|2020- | ···················· | ···················· | ···················· | ···················· | ···················· | ···················· |
| L | 2414 hCoV-19/USA/TX-DSHS-0568/2020\|EPI_ISL_574470\|2020- | ···················· | ···················· | ···················· | ···················· | ···················· | ···················· |
| L | 2415 hCoV-19/USA/TX-DSHS-0569/2020\|EPI_ISL_574471\|2020- | ···················· | ···················· | ···················· | ···················· | ···················· | ···················· |

| | | | | | | | |
|---|---|---|---|---|---|---|---|
| L | 2416 hCoV-19/USA/TX-DSHS-0570/2020\|EPI_ISL_574472\|2020- | .................... | .................... | ..................... | .................... | .................... | .................... |
| L | 2417 hCoV-19/USA/TX-DSHS-0571/2020\|EPI_ISL_574473\|2020- | .................... | .................... | ..................... | .................... | .................... | .................... |
| L | 2418 hCoV-19/USA/TX-DSHS-0572/2020\|EPI_ISL_574474\|2020- | .................... | .................... | ..................... | .................... | .................... | .................... |
| L | 2419 hCoV-19/USA/TX-DSHS-0573/2020\|EPI_ISL_574475\|2020- | .................... | .................... | ..................... | .................... | .................... | .................... |
| L | 2420 hCoV-19/USA/TX-DSHS-0574/2020\|EPI_ISL_574476\|2020- | .................... | .................... | ..................... | .................... | .................... | .................... |
| L | 2421 hCoV-19/USA/TX-DSHS-0575/2020\|EPI_ISL_574477\|2020- | .................... | .................... | ..................... | .................... | .................... | .................... |
| L | 2422 hCoV-19/USA/TX-DSHS-0576/2020\|EPI_ISL_574478\|2020- | .................... | .................... | ..................... | .................... | .................... | .................... |
| L | 2423 hCoV-19/Singapore/998/2020\|EPI_ISL_574479\|2020-03- | .................... | .................... | ..................... | .................... | .................... | .................... |
| L | 2424 hCoV-19/Singapore/52cul/2020\|EPI_ISL_574480\|2020-0 | .................... | .................... | ..................... | .................... | .................... | .................... |
| L | 2425 hCoV-19/Singapore/999/2020\|EPI_ISL_574481\|2020-03- | .................... | .................... | ..................... | .................... | .................... | .................... |
| L | 2426 hCoV-19/Singapore/1000/2020\|EPI_ISL_574482\|2020-03 | .................... | .................... | ..................... | .................... | .................... | .................... |
| L | 2427 hCoV-19/Singapore/1001/2020\|EPI_ISL_574483\|2020-03 | .................... | .................... | ..................... | .................... | .................... | .................... |
| L | 2428 hCoV-19/Singapore/141cul/2020\|EPI_ISL_574484\|2020- | .................... | .................... | ..................... | .................... | .................... | .................... |
| L | 2429 hCoV-19/Singapore/1002/2020\|EPI_ISL_574485\|2020-04 | .................... | .................... | ..................... | .................... | .................... | .................... |
| L | 2430 hCoV-19/Singapore/1003/2020\|EPI_ISL_574486\|2020-06 | .................... | .................... | ..................... | .................... | .................... | .................... |
| L | 2431 hCoV-19/Singapore/1004/2020\|EPI_ISL_574487\|2020-03 | .................... | .................... | ..................... | .................... | .................... | .................... |
| L | 2432 hCoV-19/Singapore/438cul/2020\|EPI_ISL_574488\|2020- | .................... | .................... | ..................... | .................... | .................... | .................... |
| L | 2433 hCoV-19/Singapore/1005/2020\|EPI_ISL_574489\|2020-03 | .................... | .................... | ..................... | .................... | .................... | .................... |
| L | 2434 hCoV-19/Singapore/1006/2020\|EPI_ISL_574490\|2020-03 | .................... | .................... | ..................... | .................... | .................... | .................... |
| L | 2435 hCoV-19/Singapore/1007/2020\|EPI_ISL_574491\|2020-06 | .................... | .................... | ..................... | .................... | .................... | .................... |
| L | 2436 hCoV-19/Singapore/472cul/2020\|EPI_ISL_574492\|2020- | .................... | .................... | ..................... | .................... | .................... | .................... |
| L | 2437 hCoV-19/Singapore/1008/2020\|EPI_ISL_574493\|2020-03 | .................... | .................... | ..................... | .................... | .................... | .................... |
| L | 2438 hCoV-19/Singapore/1009/2020\|EPI_ISL_574494\|2020-09 | .................... | .................... | ..................... | .................... | .................... | .................... |
| L | 2439 hCoV-19/Singapore/1010/2020\|EPI_ISL_574495\|2020-03 | .................... | .................... | ..................... | .................... | .................... | .................... |
| L | 2440 hCoV-19/Singapore/1011/2020\|EPI_ISL_574496\|2020-03 | .................... | .................... | ..................... | .................... | .................... | .................... |
| L | 2441 hCoV-19/Singapore/1012/2020\|EPI_ISL_574497\|2020-03 | .................... | .................... | ..................... | .................... | .................... | .................... |
| L | 2442 hCoV-19/Singapore/1013/2020\|EPI_ISL_574498\|2020-03 | .................... | .................... | ..................... | .................... | .................... | .................... |
| L | 2443 hCoV-19/Singapore/1014/2020\|EPI_ISL_574499\|2020-03 | .................... | .................... | ..................... | .................... | .................... | .................... |
| L | 2444 hCoV-19/England/ALDP-9CBCE6/2020\|EPI_ISL_575700\|20 | .................... | .................... | ..................... | .................... | .................... | .................... |
| L | 2445 hCoV-19/England/ALDP-9CCCC7/2020\|EPI_ISL_575701\|20 | .................... | .................... | ..................... | .................... | .................... | .................... |
| L | 2446 hCoV-19/England/ALDP-9CA9C2/2020\|EPI_ISL_575703\|20 | .................... | .................... | ..................... | .................... | .................... | .................... |
| L | 2447 hCoV-19/England/ALDP-9CB439/2020\|EPI_ISL_575704\|20 | .................... | .................... | ..................... | .................... | .................... | .................... |
| L | 2448 hCoV-19/England/ALDP-9CAC05/2020\|EPI_ISL_575705\|20 | .................... | .................... | ..................... | .................... | .................... | .................... |
| L | 2449 hCoV-19/England/ALDP-9CB32D/2020\|EPI_ISL_575706\|20 | .................... | .................... | ..................... | .................... | .................... | .................... |
| L | 2450 hCoV-19/England/ALDP-9CB8E2/2020\|EPI_ISL_575707\|20 | .................... | .................... | ..................... | .................... | .................... | .................... |
| L | 2451 hCoV-19/England/ALDP-9CBB9E/2020\|EPI_ISL_575708\|20 | .................... | .................... | ..................... | .................... | .................... | .................... |
| L | 2452 hCoV-19/England/ALDP-9CBDD4/2020\|EPI_ISL_575709\|20 | .................... | .................... | ..................... | .................... | .................... | .................... |
| L | 2453 hCoV-19/England/ALDP-9CBBF8/2020\|EPI_ISL_575720\|20 | .................... | .................... | ..................... | .................... | .................... | .................... |
| L | 2454 hCoV-19/England/ALDP-9CAC6F/2020\|EPI_ISL_575721\|20 | .................... | .A.................. | ..................... | .................... | .................... | ...A................ |
| L | 2455 hCoV-19/England/ALDP-9CAD11/2020\|EPI_ISL_575722\|20 | .................... | .................... | ..................... | .................... | .................... | .................... |
| L | 2456 hCoV-19/England/ALDP-9CB1BA/2020\|EPI_ISL_575723\|20 | .................... | .................... | ..................... | .................... | .................... | .................... |
| L | 2457 hCoV-19/England/ALDP-9CAA74/2020\|EPI_ISL_575728\|20 | .................... | .................... | ..................... | .................... | .................... | .................... |
| L | 2458 hCoV-19/England/ALDP-9CAFB1/2020\|EPI_ISL_575729\|20 | .................... | .................... | ..................... | .................... | .................... | .................... |
| L | 2459 hCoV-19/England/ALDP-9CCCB8/2020\|EPI_ISL_575730\|20 | .................... | .................... | ..................... | .................... | .................... | .................... |
| L | 2460 hCoV-19/England/MILK-97643D/2020\|EPI_ISL_575775\|20 | .................... | .................... | ..................... | .................... | .................... | .................... |
| L | 2461 hCoV-19/England/MILK-9747CD/2020\|EPI_ISL_575776\|20 | .................... | .................... | ..................... | .................... | .................... | .................... |
| L | 2462 hCoV-19/England/MILK-9736B2/2020\|EPI_ISL_575777\|20 | .................... | .................... | ..................... | .................... | .................... | .................... |
| L | 2463 hCoV-19/England/MILK-974666/2020\|EPI_ISL_575779\|20 | .................... | .................... | ..................... | .................... | .................... | .................... |
| L | 2464 hCoV-19/England/MILK-9744B7/2020\|EPI_ISL_575780\|20 | .................... | .................... | ..................... | .................... | .................... | .................... |
| L | 2465 hCoV-19/England/MILK-974718/2020\|EPI_ISL_575781\|20 | .................... | .................... | ..................... | .................... | .................... | .................... |
| L | 2466 hCoV-19/England/MILK-97352E/2020\|EPI_ISL_575782\|20 | .................... | .................... | ..................... | .................... | .................... | .................... |
| L | 2467 hCoV-19/England/MILK-9767E9/2020\|EPI_ISL_575783\|20 | .................... | .................... | ..................... | .................... | .................... | .................... |
| L | 2468 hCoV-19/England/MILK-974754/2020\|EPI_ISL_575784\|20 | .................... | .................... | ..................... | .................... | .................... | .................... |
| L | 2469 hCoV-19/England/MILK-974842/2020\|EPI_ISL_575785\|20 | .................... | .................... | ..................... | .................... | .................... | .................... |
| L | 2470 hCoV-19/England/MILK-9745E1/2020\|EPI_ISL_575786\|20 | .................... | .................... | ..................... | .................... | .................... | .................... |
| L | 2471 hCoV-19/England/MILK-9734A9/2020\|EPI_ISL_575787\|20 | .................... | .................... | ..................... | .................... | .................... | .................... |
| L | 2472 hCoV-19/England/MILK-97499A/2020\|EPI_ISL_575789\|20 | .................... | .................... | ..................... | .................... | .................... | .................... |
| L | 2473 hCoV-19/England/MILK-973290/2020\|EPI_ISL_575790\|20 | .................... | .................... | ..................... | .................... | .................... | .................... |
| L | 2474 hCoV-19/England/MILK-974A2E/2020\|EPI_ISL_575791\|20 | .................... | .................... | ..................... | .................... | .................... | .................... |
| L | 2475 hCoV-19/England/MILK-976716/2020\|EPI_ISL_575792\|20 | .................... | .................... | ..................... | .................... | .................... | .................... |
| L | 2476 hCoV-19/England/MILK-97644C/2020\|EPI_ISL_575793\|20 | .................... | .................... | ..................... | .................... | .................... | .................... |
| L | 2477 hCoV-19/England/MILK-974903/2020\|EPI_ISL_575794\|20 | .................... | .................... | ..................... | .................... | .................... | .................... |
| L | 2478 hCoV-19/England/MILK-974727/2020\|EPI_ISL_575795\|20 | .................... | .................... | ..................... | .................... | .................... | .................... |
| L | 2479 hCoV-19/England/MILK-9748E8/2020\|EPI_ISL_575797\|20 | .................... | .................... | ..................... | .................... | .................... | .................... |
| L | 2480 hCoV-19/England/MILK-97344F/2020\|EPI_ISL_575798\|20 | .................... | .................... | ..................... | .................... | .................... | .................... |
| L | 2481 hCoV-19/England/MILK-9764F1/2020\|EPI_ISL_575799\|20 | .................... | .A.................. | ..................... | .................... | .................... | .................... |
| L | 2483 hCoV-19/New Zealand/20VR2007/2020\|EPI_ISL_579201\|2 | .................... | .................... | ..................... | .................... | .................... | .................... |
| L | 2484 hCoV-19/New Zealand/20VR2009/2020\|EPI_ISL_579202\|2 | .................... | .................... | ..................... | .................... | .................... | .................... |
| L | 2487 hCoV-19/New Zealand/20VR2013/2020\|EPI_ISL_579205\|2 | .................... | .................... | ..................... | .................... | .................... | .................... |
| L | 2488 hCoV-19/New Zealand/20VR2016/2020\|EPI_ISL_579206\|2 | .................... | .................... | ..................... | .................... | .................... | .................... |
| L | 2489 hCoV-19/New Zealand/20VR2018/2020\|EPI_ISL_579207\|2 | .................... | .................... | ..................... | .................... | .................... | .................... |
| L | 2490 hCoV-19/New Zealand/20VR2021/2020\|EPI_ISL_579209\|2 | .................... | .................... | ..................... | .................... | .................... | .................... |
| L | 2491 hCoV-19/New Zealand/20VR2023/2020\|EPI_ISL_579210\|2 | .................... | .................... | ..................... | .................... | .................... | .................... |
| L | 2492 hCoV-19/New Zealand/20VR2026/2020\|EPI_ISL_579213\|2 | .................... | .................... | ..................... | .................... | .................... | .................... |
| L | 2493 hCoV-19/New Zealand/20VR2027/2020\|EPI_ISL_579214\|2 | .................... | .................... | ..................... | .................... | .................... | .................... |
| L | 2494 hCoV-19/New Zealand/20VR2029/2020\|EPI_ISL_579215\|2 | .................... | .................... | ..................... | .................... | .................... | .................... |
| L | 2495 hCoV-19/New Zealand/20VR2031/2020\|EPI_ISL_579216\|2 | .................... | .................... | ..................... | .................... | .................... | .................... |
| L | 2496 hCoV-19/New Zealand/20VR2036/2020\|EPI_ISL_579217\|2 | .................... | .................... | ..................... | .................... | .................... | .................... |
| L | 2497 hCoV-19/England/CAMC-9EC54E/2020\|EPI_ISL_580286\|20 | .................... | .................... | ..................... | .................... | .................... | .................... |
| L | 2498 hCoV-19/England/CAMC-9EC0C5/2020\|EPI_ISL_580287\|20 | .................... | .................... | ..................... | .................... | .................... | .................... |

| | | | | | | | | |
|---|---|---|---|---|---|---|---|---|
| L | 2499 | hCoV-19/England/ALDP-9E608F/2020\|EPI_ISL_580288\|20 | ···················· | ·A·······A·········· | ···················· | ···················· | ···················· | ···················· |
| L | 2500 | hCoV-19/England/ALDP-9E6D88/2020\|EPI_ISL_580289\|20 | ···················· | ···················· | ···················· | ···················· | ···················· | ···················· |
| L | 2501 | hCoV-19/England/ALDP-9E60CB/2020\|EPI_ISL_580292\|20 | ···················· | ···················· | ···················· | ···················· | ···················· | ···················· |
| L | 2502 | hCoV-19/England/ALDP-9E7394/2020\|EPI_ISL_580293\|20 | ···················· | ···················· | ···················· | ···················· | ···················· | ···················· |
| L | 2503 | hCoV-19/England/ALDP-9E692A/2020\|EPI_ISL_580294\|20 | ···················· | ···················· | ···················· | ···················· | ···················· | ···················· |
| L | 2504 | hCoV-19/England/ALDP-9E6614/2020\|EPI_ISL_580295\|20 | ···················· | ···················· | ···················· | ···················· | ···················· | ···················· |
| L | 2505 | hCoV-19/India/WB-IICB-025/2020\|EPI_ISL_581500\|2020 | ···················· | ···················· | ···················· | ···················· | ···················· | ···················· |
| L | 2506 | hCoV-19/India/WB-IICB-026/2020\|EPI_ISL_581501\|2020 | ···················· | ···················· | ···················· | ···················· | ···················· | ···················· |
| L | 2507 | hCoV-19/India/WB-IICB-027/2020\|EPI_ISL_581502\|2020 | ···················· | ···················· | ···················· | ···················· | ···················· | ···················· |
| L | 2508 | hCoV-19/India/WB-IICB-028/2020\|EPI_ISL_581503\|2020 | ···················· | ···················· | ···················· | ···················· | ···················· | ···················· |
| L | 2509 | hCoV-19/India/WB-IICB-029/2020\|EPI_ISL_581504\|2020 | ···················· | ···················· | ···················· | ···················· | ···················· | ···················· |
| L | 2510 | hCoV-19/India/DL-NCDC-IGIB-77797A/2020\|EPI_ISL_581 | ···················· | ·A·················· | ···················· | ···················· | ···················· | ···A················ |
| L | 2511 | hCoV-19/India/DL-NCDC-IGIB-D1/2020\|EPI_ISL_581507\| | ···················· | ·A·················· | ···················· | ···················· | ···················· | ···A················ |
| L | 2512 | hCoV-19/USA/VA-DCLS-1506/2020\|EPI_ISL_581508\|2020- | ···················· | ···················· | ···················· | ···················· | ···················· | ···················· |
| L | 2513 | hCoV-19/USA/VA-DCLS-1507/2020\|EPI_ISL_581509\|2020- | ···················· | ···················· | ···················· | ···················· | ···················· | ···················· |
| L | 2514 | hCoV-19/USA/VA-DCLS-1508/2020\|EPI_ISL_581510\|2020- | ···················· | ···················· | ···················· | ···················· | ···················· | ···················· |
| L | 2515 | hCoV-19/USA/VA-DCLS-1512/2020\|EPI_ISL_581511\|2020- | ···················· | ···················· | ···················· | ···················· | ···················· | ···················· |
| L | 2516 | hCoV-19/USA/VA-DCLS-1513/2020\|EPI_ISL_581512\|2020- | ···················· | ···················· | ···················· | ···················· | ···················· | ···················· |
| L | 2517 | hCoV-19/USA/VA-DCLS-1514/2020\|EPI_ISL_581513\|2020- | ···················· | ···················· | ···················· | ···················· | ···················· | ···················· |
| L | 2518 | hCoV-19/USA/VA-DCLS-1517/2020\|EPI_ISL_581514\|2020- | ···················· | ···················· | ···················· | ···················· | ···················· | ···················· |
| L | 2519 | hCoV-19/USA/VA-DCLS-1518/2020\|EPI_ISL_581515\|2020- | ···················· | ···················· | ···················· | ···················· | ···················· | ···················· |
| L | 2520 | hCoV-19/USA/VA-DCLS-1519/2020\|EPI_ISL_581516\|2020- | ···················· | ···················· | ···················· | ···················· | ···················· | ···················· |
| L | 2521 | hCoV-19/USA/VA-DCLS-1520/2020\|EPI_ISL_581517\|2020- | ···················· | ···················· | ···················· | ···················· | ···················· | ···················· |
| L | 2522 | hCoV-19/USA/VA-DCLS-1521/2020\|EPI_ISL_581518\|2020- | ···················· | ···················· | ···················· | ···················· | ···················· | ···················· |
| L | 2523 | hCoV-19/USA/VA-DCLS-1523/2020\|EPI_ISL_581520\|2020- | ···················· | ···················· | ···················· | ···················· | ···················· | ···················· |
| L | 2524 | hCoV-19/USA/VA-DCLS-1527/2020\|EPI_ISL_581523\|2020- | ···················· | ···················· | ···················· | ···················· | ···················· | ···················· |
| L | 2525 | hCoV-19/USA/VA-DCLS-1528/2020\|EPI_ISL_581524\|2020- | ···················· | ···················· | ···················· | ···················· | ···················· | ···················· |
| L | 2526 | hCoV-19/USA/VA-DCLS-1529/2020\|EPI_ISL_581525\|2020- | ···················· | ·············A······ | ···················· | ···················· | ···················· | ···················· |
| L | 2527 | hCoV-19/USA/VA-DCLS-1530/2020\|EPI_ISL_581526\|2020- | ···················· | ·A·················· | ···················· | ···················· | ···················· | ···················· |
| L | 2528 | hCoV-19/USA/VA-DCLS-1531/2020\|EPI_ISL_581527\|2020- | ···················· | ···················· | ···················· | ···················· | ···················· | ···················· |
| L | 2529 | hCoV-19/USA/VA-DCLS-1532/2020\|EPI_ISL_581528\|2020- | ···················· | ·A·················· | ···················· | ···················· | ···················· | ···················· |
| L | 2530 | hCoV-19/USA/VA-DCLS-1533/2020\|EPI_ISL_581529\|2020- | ···················· | ···················· | ···················· | ···················· | ···················· | ···················· |
| L | 2531 | hCoV-19/Spain/MD-IBV-99007326/2020\|EPI_ISL_582814\| | ···················· | ···················· | ···················· | ···················· | ···················· | ···················· |
| L | 2534 | hCoV-19/Italy/SIC-IZSBM169-64/2020\|EPI_ISL_582833\| | ···················· | ···················· | ···················· | ···················· | ···················· | ···················· |
| L | 2535 | hCoV-19/Sweden/20-51886/2020\|EPI_ISL_582834\|2020-0 | ···················· | ···················· | ···················· | ···················· | ···················· | ···················· |
| L | 2536 | hCoV-19/Sweden/20-52115/2020\|EPI_ISL_582835\|2020-0 | ···················· | ···················· | ···················· | ···················· | ···················· | ···················· |
| L | 2537 | hCoV-19/Sweden/20-52182/2020\|EPI_ISL_582836\|2020-0 | ···················· | ···················· | ···················· | ···················· | ···················· | ···················· |
| L | 2538 | hCoV-19/Sweden/20-52184/2020\|EPI_ISL_582837\|2020-0 | ···················· | ···················· | ···················· | ···················· | ···················· | ···················· |
| L | 2539 | hCoV-19/Sweden/20-52185/2020\|EPI_ISL_582838\|2020-0 | ···················· | ···················· | ···················· | ···················· | ···················· | ···················· |
| L | 2540 | hCoV-19/Sweden/20-52645/2020\|EPI_ISL_582839\|2020-0 | ···················· | ···················· | ···················· | ···················· | ······T············· | ···················· |
| L | 2541 | hCoV-19/Sweden/20-52777/2020\|EPI_ISL_582840\|2020-0 | ···················· | ····A··············· | ···················· | ···················· | ···················· | ···················· |
| L | 2542 | hCoV-19/Sweden/20-52943/2020\|EPI_ISL_582841\|2020-0 | ···················· | ···················· | ···················· | ···················· | ···················· | ···················· |
| L | 2543 | hCoV-19/Italy/SIC-IZSBM193-88/2020\|EPI_ISL_582842\| | ···················· | ···················· | ···················· | ···················· | ···················· | ···················· |
| L | 2544 | hCoV-19/Spain/MD-IBV-99007733/2020\|EPI_ISL_582843\| | ···················· | ···················· | ···················· | ···················· | ···················· | ···················· |
| L | 2545 | hCoV-19/Spain/MD-IBV-99007734/2020\|EPI_ISL_582844\| | ···················· | ···················· | ···················· | ···················· | ···················· | ···················· |
| L | 2546 | hCoV-19/Italy/SIC-IZSBM193-69/2020\|EPI_ISL_582845\| | ···················· | ···················· | ···················· | ···················· | ···················· | ···················· |
| L | 2547 | hCoV-19/Italy/SIC-IZSBM193-67/2020\|EPI_ISL_582846\| | ···················· | ···················· | ···················· | ···················· | ···················· | ···················· |
| L | 2548 | hCoV-19/Italy/SIC-IZSBM194-48/2020\|EPI_ISL_582847\| | ···················· | ···················· | ···················· | ···················· | ···················· | ···················· |
| L | 2549 | hCoV-19/Italy/SIC-IZSBM194-11/2020\|EPI_ISL_582848\| | ···················· | ···················· | ···················· | ···················· | ···················· | ···················· |
| L | 2550 | hCoV-19/Italy/SIC-IZSBM193-72/2020\|EPI_ISL_582849\| | ···················· | ···················· | ···················· | ···················· | ···················· | ···················· |
| L | 2551 | hCoV-19/Italy/SIC-IZSBM193-85/2020\|EPI_ISL_582850\| | ···················· | ···················· | ···················· | ···················· | ···················· | ···················· |
| L | 2552 | hCoV-19/USA/CA-CZB-2743/2020\|EPI_ISL_582851\|2020-0 | ···················· | ·A·················· | ···················· | ···················· | ···················· | ···················· |
| L | 2553 | hCoV-19/USA/CA-CZB-2744/2020\|EPI_ISL_582852\|2020-0 | ···················· | ·A·················· | ···················· | ···················· | ···················· | ···················· |
| L | 2554 | hCoV-19/USA/CA-CZB-2745/2020\|EPI_ISL_582853\|2020-0 | ···················· | ·A·················· | ···················· | ···················· | ···················· | ···················· |
| L | 2555 | hCoV-19/USA/CA-CZB-2746/2020\|EPI_ISL_582854\|2020-0 | ···················· | ·A·················· | ···················· | ···················· | ···················· | ···················· |
| L | 2556 | hCoV-19/USA/CA-CZB-2747/2020\|EPI_ISL_582855\|2020-0 | ···················· | ·A·················· | ···················· | ···················· | ···················· | ···················· |
| L | 2557 | hCoV-19/USA/CA-CZB-2748/2020\|EPI_ISL_582856\|2020-0 | ···················· | ·A·················· | ···················· | ···················· | ···················· | ···················· |
| L | 2558 | hCoV-19/USA/CA-CZB-2750/2020\|EPI_ISL_582857\|2020-0 | ···················· | ·A·················· | ···················· | ···················· | ···················· | ···················· |
| L | 2559 | hCoV-19/USA/CA-CZB-2751/2020\|EPI_ISL_582858\|2020-0 | ···················· | ·A·················· | ···················· | ···················· | ···················· | ···················· |
| L | 2560 | hCoV-19/USA/CA-CZB-2753/2020\|EPI_ISL_582859\|2020-0 | ···················· | ·A·················· | ···················· | ···················· | ···················· | ···················· |
| L | 2561 | hCoV-19/England/ALDP-9CC33B/2020\|EPI_ISL_573100\|20 | ···················· | ······C············· | ···················· | ···················· | ···················· | ···················· |
| L | 2562 | hCoV-19/England/ALDP-9CC368/2020\|EPI_ISL_573101\|20 | ···················· | ···················· | ···················· | ···················· | ···················· | ···················· |
| L | 2563 | hCoV-19/England/ALDP-9CC395/2020\|EPI_ISL_573102\|20 | ···················· | ·A·················· | ···················· | ···················· | ···················· | ···A················ |
| L | 2564 | hCoV-19/England/ALDP-9CC3A4/2020\|EPI_ISL_573103\|20 | ···················· | ···················· | ···················· | ···················· | ···················· | ···················· |
| L | 2571 | hCoV-19/England/ALDP-9CC4B0/2020\|EPI_ISL_573110\|20 | ···················· | ···················· | ···················· | ···················· | ···················· | ···················· |
| L | 2572 | hCoV-19/England/ALDP-9CC4ED/2020\|EPI_ISL_573111\|20 | ···················· | ···················· | ···················· | ···················· | ···················· | ···················· |
| L | 2574 | hCoV-19/USA/LA-EVTL266/2020\|EPI_ISL_574400\|2020-08 | ···················· | ···················· | ···················· | ···················· | ···················· | ···················· |
| L | 2575 | hCoV-19/USA/LA-EVTL267/2020\|EPI_ISL_574401\|2020-08 | ···················· | ···················· | ···················· | ···················· | ···················· | ···················· |
| L | 2576 | hCoV-19/USA/LA-EVTL268/2020\|EPI_ISL_574402\|2020-08 | ···················· | ···················· | ···················· | ···················· | ···················· | ···················· |
| L | 2577 | hCoV-19/USA/LA-EVTL269/2020\|EPI_ISL_574403\|2020-08 | ···················· | ···················· | ···················· | ···················· | ···················· | ···················· |
| L | 2578 | hCoV-19/USA/LA-EVTL270/2020\|EPI_ISL_574404\|2020-08 | ···················· | ···················· | ···················· | ···················· | ···················· | ···················· |
| L | 2579 | hCoV-19/USA/LA-EVTL271/2020\|EPI_ISL_574405\|2020-08 | ···················· | ···················· | ···················· | ···················· | ···················· | ···················· |
| L | 2580 | hCoV-19/USA/LA-EVTL272/2020\|EPI_ISL_574406\|2020-08 | ···················· | ···················· | ···················· | ···················· | ···················· | ···················· |
| L | 2581 | hCoV-19/USA/LA-EVTL273/2020\|EPI_ISL_574407\|2020-08 | ···················· | ···················· | ···················· | ···················· | ···················· | ···················· |
| L | 2582 | hCoV-19/USA/LA-EVTL274/2020\|EPI_ISL_574408\|2020-08 | ···················· | ···················· | ···················· | ···················· | ···················· | ···················· |
| L | 2583 | hCoV-19/USA/LA-EVTL275/2020\|EPI_ISL_574409\|2020-08 | ···················· | ···················· | ···················· | ···················· | ···················· | ···················· |
| L | 2584 | hCoV-19/USA/LA-EVTL276/2020\|EPI_ISL_574410\|2020-08 | ···················· | ···················· | ···················· | ···················· | ···················· | ···················· |
| L | 2585 | hCoV-19/USA/LA-EVTL277/2020\|EPI_ISL_574411\|2020-08 | ···················· | ···················· | ···················· | ···················· | ···················· | ···················· |
| L | 2586 | hCoV-19/USA/LA-EVTL278/2020\|EPI_ISL_574412\|2020-08 | ···················· | ···················· | ···················· | ···················· | ···················· | ···················· |
| L | 2587 | hCoV-19/USA/LA-EVTL279/2020\|EPI_ISL_574413\|2020-08 | ···················· | ···················· | ···················· | ···················· | ···················· | ···················· |

| | | | | | | | |
|---|---|---|---|---|---|---|---|
| L | 2588 hCoV-19/USA/LA-EVTL280/2020\|EPI_ISL_574414\|2020-08 | ···················· | ···················· | ···················· | ···················· | ···················· | ···················· |
| L | 2589 hCoV-19/USA/LA-EVTL281/2020\|EPI_ISL_574415\|2020-08 | ···················· | ···················· | ···················· | ···················· | ···················· | ···················· |
| L | 2590 hCoV-19/USA/LA-EVTL282/2020\|EPI_ISL_574416\|2020-08 | ···················· | ···················· | ···················· | ···················· | ···················· | ···················· |
| L | 2591 hCoV-19/USA/LA-EVTL283/2020\|EPI_ISL_574417\|2020-08 | ···················· | ···················· | ···················· | ···················· | ···················· | ···················· |
| L | 2592 hCoV-19/USA/LA-EVTL284/2020\|EPI_ISL_574418\|2020-08 | ···················· | ···················· | ···················· | ···················· | ···················· | ···················· |
| L | 2593 hCoV-19/USA/LA-EVTL285/2020\|EPI_ISL_574419\|2020-08 | ···················· | ···················· | ···················· | ···················· | ···················· | ···················· |
| L | 2594 hCoV-19/USA/LA-EVTL286/2020\|EPI_ISL_574420\|2020-08 | ···················· | ·A·················· | ···················· | ···················· | ···················· | ···················· |
| L | 2595 hCoV-19/USA/LA-EVTL287/2020\|EPI_ISL_574421\|2020-08 | ···················· | ···················· | ···················· | ···················· | ···················· | ···················· |
| L | 2596 hCoV-19/USA/LA-EVTL288/2020\|EPI_ISL_574422\|2020-08 | ···················· | ···················· | ···················· | ···················· | ···················· | ···················· |
| L | 2597 hCoV-19/USA/LA-EVTL289/2020\|EPI_ISL_574423\|2020-08 | ···················· | ···················· | ···················· | ···················· | ···················· | ···················· |
| L | 2598 hCoV-19/USA/LA-EVTL290/2020\|EPI_ISL_574424\|2020-08 | ···················· | ···················· | ···················· | ···················· | ···················· | ···················· |
| L | 2599 hCoV-19/USA/LA-EVTL291/2020\|EPI_ISL_574425\|2020-08 | ···················· | ·A·················· | ···················· | ···················· | ···················· | ···················· |
| L | 2600 hCoV-19/USA/LA-EVTL292/2020\|EPI_ISL_574426\|2020-08 | ···················· | ···················· | ···················· | ···················· | ···················· | ···················· |
| L | 2601 hCoV-19/USA/LA-EVTL293/2020\|EPI_ISL_574427\|2020-08 | ···················· | ···················· | ···················· | ···················· | ···················· | ···················· |
| L | 2602 hCoV-19/USA/LA-EVTL294/2020\|EPI_ISL_574428\|2020-08 | ···················· | ···················· | ···················· | ···················· | ···················· | ···················· |
| L | 2603 hCoV-19/USA/LA-EVTL295/2020\|EPI_ISL_574429\|2020-08 | ···················· | ···················· | ···················· | ···················· | ···················· | ···················· |
| L | 2604 hCoV-19/USA/LA-EVTL296/2020\|EPI_ISL_574430\|2020-08 | ···················· | ···················· | ···················· | ···················· | ···················· | ···················· |
| L | 2605 hCoV-19/Ecuador/USFQ-253/2020\|EPI_ISL_574431\|2020- | ···················· | ···················· | ···················· | ···················· | ···················· | ···················· |
| L | 2606 hCoV-19/Indonesia/JK-NIHRD-C0027475/2020\|EPI_ISL_5 | ···················· | ···A················ | ···················· | ···················· | ···················· | ···················· |
| L | 2607 hCoV-19/USA/TX-DSHS-0530/2020\|EPI_ISL_574435\|2020- | ···················· | ···················· | ···················· | ···················· | ···················· | ···················· |
| L | 2608 hCoV-19/USA/TX-DSHS-0531/2020\|EPI_ISL_574436\|2020- | ···················· | ···················· | ···················· | ···················· | ···················· | ···················· |
| L | 2609 hCoV-19/USA/TX-DSHS-0533/2020\|EPI_ISL_574438\|2020- | ···················· | ···················· | ···················· | ···················· | ···················· | ···················· |
| L | 2610 hCoV-19/USA/TX-DSHS-0534/2020\|EPI_ISL_574439\|2020- | ···················· | ···················· | ···················· | ···················· | ···················· | ···················· |
| L | 2611 hCoV-19/USA/TX-DSHS-0535/2020\|EPI_ISL_574440\|2020- | ···················· | ···················· | ···················· | ···················· | ···················· | ···················· |
| L | 2612 hCoV-19/USA/TX-DSHS-0536/2020\|EPI_ISL_574441\|2020- | ···················· | ···················· | ···················· | ···················· | ···················· | ···················· |
| L | 2613 hCoV-19/USA/TX-DSHS-0539/2020\|EPI_ISL_574443\|2020- | ···················· | ···················· | ···················· | ···················· | ···················· | ···················· |
| L | 2614 hCoV-19/England/ALDP-9CAB26/2020\|EPI_ISL_575710\|20 | ···················· | ···················· | ···················· | ···················· | ···················· | ···················· |
| L | 2615 hCoV-19/England/ALDP-9CB5FA/2020\|EPI_ISL_575711\|20 | ···················· | ···················· | ···················· | ···················· | ···················· | ···················· |
| L | 2616 hCoV-19/England/ALDP-9CB008/2020\|EPI_ISL_575712\|20 | ···················· | ···················· | ···················· | ···················· | ···················· | ···················· |
| L | 2617 hCoV-19/England/ALDP-9CB703/2020\|EPI_ISL_575713\|20 | ···················· | ···················· | ···················· | ···················· | ···················· | ···················· |
| L | 2618 hCoV-19/England/ALDP-9CACF6/2020\|EPI_ISL_575715\|20 | ···················· | ···················· | ···················· | ···················· | ···················· | ···················· |
| L | 2619 hCoV-19/England/ALDP-9CB5AF/2020\|EPI_ISL_575716\|20 | ···················· | ···················· | ···················· | ···················· | ···················· | ···················· |
| L | 2620 hCoV-19/England/ALDP-9CAE4B/2020\|EPI_ISL_575717\|20 | ···················· | ···················· | ···················· | ···················· | ···················· | ···················· |
| L | 2621 hCoV-19/England/ALDP-9CAF1B/2020\|EPI_ISL_575719\|20 | ···················· | ···················· | ···················· | ···················· | ···················· | ···················· |
| L | 2622 hCoV-19/England/ALDP-9CB624/2020\|EPI_ISL_575724\|20 | ···················· | ···················· | ···················· | ···················· | ···················· | ···················· |
| L | 2623 hCoV-19/England/ALDP-9CAFEE/2020\|EPI_ISL_575725\|20 | ···················· | ···················· | ···················· | ···················· | ···················· | ···················· |
| L | 2624 hCoV-19/England/ALDP-9CB220/2020\|EPI_ISL_575726\|20 | ···················· | ···················· | ···················· | ···················· | ···················· | ···················· |
| L | 2625 hCoV-19/England/ALDP-9CACBA/2020\|EPI_ISL_575727\|20 | ···················· | ···················· | ···················· | ···················· | ···················· | ···················· |
| L | 2626 hCoV-19/England/ALDP-9CB3B4/2020\|EPI_ISL_575731\|20 | ···················· | ···················· | ···················· | ···················· | ···················· | ···················· |
| L | 2627 hCoV-19/England/ALDP-9CB8B5/2020\|EPI_ISL_575732\|20 | ···················· | ···················· | ···················· | ···················· | ···················· | ···················· |
| L | 2628 hCoV-19/England/ALDP-9CB044/2020\|EPI_ISL_575733\|20 | ···················· | ···················· | ···················· | ···················· | ···················· | ···················· |
| L | 2629 hCoV-19/England/ALDP-9CAA92/2020\|EPI_ISL_575734\|20 | ···················· | ···················· | ···················· | ···················· | ···················· | ···················· |
| L | 2630 hCoV-19/England/ALDP-9CB4C0/2020\|EPI_ISL_575736\|20 | ···················· | ···················· | ···················· | ···················· | ···················· | ···················· |
| L | 2631 hCoV-19/England/ALDP-9CB3A5/2020\|EPI_ISL_575737\|20 | ···················· | ···················· | ···················· | ···················· | ···················· | ···················· |
| L | 2632 hCoV-19/England/ALDP-9CB035/2020\|EPI_ISL_575738\|20 | ···················· | ···················· | ···················· | ···················· | ···················· | ···················· |
| L | 2633 hCoV-19/England/ALDP-9CAF93/2020\|EPI_ISL_575739\|20 | ···················· | ···················· | ···················· | ···················· | ···················· | ···················· |
| L | 2634 hCoV-19/England/ALDP-9CAD20/2020\|EPI_ISL_575740\|20 | ···················· | ·A·················· | ···················· | ···················· | ···················· | ··A················· |
| L | 2635 hCoV-19/England/ALDP-9CADF3/2020\|EPI_ISL_575741\|20 | ···················· | ·A·················· | ···················· | ···················· | ···················· | ··A················· |
| L | 2636 hCoV-19/England/ALDP-9CBA0A/2020\|EPI_ISL_575743\|20 | ···················· | ···················· | ···················· | ···················· | ···················· | ···················· |
| L | 2637 hCoV-19/England/ALDP-9CABF9/2020\|EPI_ISL_575744\|20 | ···················· | ·A·················· | ···················· | ···················· | ···················· | ··A················· |
| L | 2638 hCoV-19/England/ALDP-9CBABF/2020\|EPI_ISL_575745\|20 | ···················· | ···················· | ···················· | ···················· | ···················· | ···················· |
| L | 2639 hCoV-19/England/ALDP-9CB0DB/2020\|EPI_ISL_575746\|20 | ···················· | ···················· | ···················· | ···················· | ···················· | ···················· |
| L | 2640 hCoV-19/England/ALDP-9CB1D8/2020\|EPI_ISL_575747\|20 | ···················· | ·A·················· | ···················· | ···················· | ···················· | ··A················· |
| L | 2641 hCoV-19/England/ALDP-9CAEB4/2020\|EPI_ISL_575748\|20 | ···················· | ···················· | ···················· | ···················· | ···················· | ···················· |
| L | 2642 hCoV-19/England/ALDP-9CBCB9/2020\|EPI_ISL_575749\|20 | ···················· | ···················· | ···················· | ···················· | ···················· | ···················· |
| L | 2643 hCoV-19/England/ALDP-9CAB62/2020\|EPI_ISL_575751\|20 | ···················· | ···················· | ···················· | ···················· | ···················· | ···················· |
| L | 2644 hCoV-19/England/ALDP-9CAE2D/2020\|EPI_ISL_575754\|20 | ···················· | ···················· | ···················· | ···················· | ···················· | ···················· |
| L | 2645 hCoV-19/England/ALDP-9CAAED/2020\|EPI_ISL_575756\|20 | ···················· | ···················· | ···················· | ···················· | ···················· | ···················· |
| L | 2646 hCoV-19/England/ALDP-9CB967/2020\|EPI_ISL_575757\|20 | ···················· | ···················· | ···················· | ···················· | ···················· | ···················· |
| L | 2647 hCoV-19/England/ALDP-9CB448/2020\|EPI_ISL_575758\|20 | ···················· | ···················· | ···················· | ···················· | ···················· | ···················· |
| L | 2648 hCoV-19/England/ALDP-9CC016/2020\|EPI_ISL_575760\|20 | ···················· | ·A·················· | ···················· | ···················· | ···················· | ···················· |
| L | 2649 hCoV-19/England/MILK-9765D0/2020\|EPI_ISL_575762\|20 | ···················· | ···················· | ···················· | ···················· | ···················· | ···················· |
| L | 2650 hCoV-19/Wales/MILK-973139/2020\|EPI_ISL_575763\|2020 | ···················· | ···················· | ···················· | ···················· | ···················· | ···················· |
| L | 2651 hCoV-19/England/MILK-9749E5/2020\|EPI_ISL_575764\|20 | ···················· | ···················· | ···················· | ···················· | ···················· | ···················· |
| L | 2652 hCoV-19/England/MILK-974A6A/2020\|EPI_ISL_575765\|20 | ···················· | ···················· | ···················· | ···················· | ···················· | ···················· |
| L | 2653 hCoV-19/England/MILK-974684/2020\|EPI_ISL_575766\|20 | ···················· | ···················· | ···················· | ···················· | ···················· | ···················· |
| L | 2654 hCoV-19/England/MILK-9731A2/2020\|EPI_ISL_575768\|20 | ···················· | ···················· | ···················· | ···················· | ···················· | ···················· |
| L | 2655 hCoV-19/England/MILK-974596/2020\|EPI_ISL_575769\|20 | ···················· | ···················· | ···················· | ···················· | ···················· | ···················· |
| L | 2656 hCoV-19/England/MILK-9765EF/2020\|EPI_ISL_575770\|20 | ···················· | ···················· | ···················· | ···················· | ···················· | ···················· |
| L | 2657 hCoV-19/Wales/MILK-973588/2020\|EPI_ISL_575771\|2020 | ···················· | ···················· | ···················· | ···················· | ···················· | ···················· |
| L | 2658 hCoV-19/England/MILK-974824/2020\|EPI_ISL_575772\|20 | ···················· | ···················· | ···················· | ···················· | ···················· | ···················· |
| L | 2659 hCoV-19/England/MILK-974B58/2020\|EPI_ISL_575773\|20 | ···················· | ···················· | ···················· | ···················· | ···················· | ···················· |
| L | 2660 hCoV-19/England/MILK-9766DD/2020\|EPI_ISL_575774\|20 | ···················· | ···················· | ···················· | ···················· | ···················· | ···················· |
| L | 2661 hCoV-19/New Zealand/20CV0354/2020\|EPI_ISL_579100\|2 | ···················· | ·A·················· | ···················· | ···················· | ···················· | ··A················· |
| L | 2662 hCoV-19/New Zealand/20CV0356/2020\|EPI_ISL_579102\|2 | ···················· | ···················· | ···················· | ···················· | ···················· | ···················· |
| L | 2663 hCoV-19/New Zealand/20CV0357/2020\|EPI_ISL_579103\|2 | ···················· | ·A·················· | ···················· | ···················· | ···················· | ··A················· |
| L | 2664 hCoV-19/New Zealand/20CV0358/2020\|EPI_ISL_579104\|2 | ···················· | ·A·················· | ···················· | ···················· | ···················· | ··A················· |
| L | 2665 hCoV-19/New Zealand/20CV0362/2020\|EPI_ISL_579105\|2 | ···················· | ·A·················· | ···················· | ···················· | ···················· | ··A················· |
| L | 2666 hCoV-19/New Zealand/20CV0363/2020\|EPI_ISL_579106\|2 | ···················· | ···················· | ···················· | ···················· | ···················· | ···················· |
| L | 2667 hCoV-19/New Zealand/20CV0364/2020\|EPI_ISL_579107\|2 | ···················· | ·A·················· | ···················· | ···················· | ···················· | ··A················· |

| | | | | | | | |
|---|---|---|---|---|---|---|---|
| L | 2668 hCoV-19/New Zealand/20CV0366/2020\|EPI_ISL_579109\|2 | ···················· | ···················· | ···················· | ···················· | ······T············· | ···················· |
| L | 2669 hCoV-19/New Zealand/20CV0369/2020\|EPI_ISL_579111\|2 | ···················· | ···················· | ···················· | ···················· | ···················· | ···················· |
| L | 2670 hCoV-19/New Zealand/20CV0375/2020\|EPI_ISL_579113\|2 | ···················· | ·A·················· | ···················· | ···················· | ···················· | ···A················ |
| L | 2671 hCoV-19/New Zealand/20CV0380/2020\|EPI_ISL_579115\|2 | ···················· | ·A·················· | ···················· | ···················· | ···················· | ···A················ |
| L | 2672 hCoV-19/New Zealand/20CV0387/2020\|EPI_ISL_579116\|2 | ···················· | ···················· | ···················· | ···················· | ···················· | ···················· |
| L | 2673 hCoV-19/New Zealand/20VR0769/2020\|EPI_ISL_579118\|2 | ···················· | ···················· | ···················· | ···················· | ···················· | ···················· |
| L | 2674 hCoV-19/New Zealand/20VR0997/2020\|EPI_ISL_579119\|2 | ···················· | ···················· | ···················· | ···················· | ···················· | ···················· |
| L | 2675 hCoV-19/New Zealand/20VR1113/2020\|EPI_ISL_579121\|2 | ···················· | ···················· | ···················· | ···················· | ···················· | ···················· |
| L | 2676 hCoV-19/New Zealand/20VR1291/2020\|EPI_ISL_579122\|2 | ···················· | ···················· | ···················· | ···················· | ···················· | ···················· |
| L | 2679 hCoV-19/New Zealand/20VR1764/2020\|EPI_ISL_579126\|2 | ···················· | ···················· | ···················· | ···················· | ···················· | ···················· |
| L | 2681 hCoV-19/New Zealand/20VR1775/2020\|EPI_ISL_579128\|2 | ···················· | ···················· | ···················· | ···················· | ···················· | ···················· |
| L | 2682 hCoV-19/New Zealand/20VR1792/2020\|EPI_ISL_579129\|2 | ···················· | ···················· | ···················· | ···················· | ···················· | ···················· |
| L | 2683 hCoV-19/New Zealand/20VR1793/2020\|EPI_ISL_579130\|2 | ···················· | ···················· | ···················· | ···················· | ···················· | ···················· |
| L | 2684 hCoV-19/New Zealand/20VR1794/2020\|EPI_ISL_579131\|2 | ···················· | ···················· | ···················· | ···················· | ···················· | ···················· |
| L | 2685 hCoV-19/New Zealand/20VR1795/2020\|EPI_ISL_579132\|2 | ···················· | ···················· | ···················· | ···················· | ···················· | ···················· |
| L | 2686 hCoV-19/New Zealand/20VR1806/2020\|EPI_ISL_579133\|2 | ···················· | ···················· | ···················· | ···················· | ···················· | ···················· |
| L | 2687 hCoV-19/New Zealand/20VR1807/2020\|EPI_ISL_579134\|2 | ···················· | ···················· | ···················· | ···················· | ···················· | ···················· |
| L | 2688 hCoV-19/New Zealand/20VR1808/2020\|EPI_ISL_579135\|2 | ···················· | ···················· | ···················· | ···················· | ···················· | ···················· |
| L | 2689 hCoV-19/New Zealand/20VR1811/2020\|EPI_ISL_579136\|2 | ···················· | ···················· | ···················· | ···················· | ···················· | ···················· |
| L | 2690 hCoV-19/New Zealand/20VR1813/2020\|EPI_ISL_579138\|2 | ···················· | ·A·················· | ···················· | ···················· | ···················· | ···················· |
| L | 2691 hCoV-19/New Zealand/20VR1815/2020\|EPI_ISL_579139\|2 | ···················· | ·A·················· | ···················· | ···················· | ···················· | ···················· |
| L | 2692 hCoV-19/New Zealand/20VR1819/2020\|EPI_ISL_579140\|2 | ···················· | ···················· | ···················· | ···················· | ···················· | ···················· |
| L | 2693 hCoV-19/New Zealand/20VR1820/2020\|EPI_ISL_579141\|2 | ···················· | ···················· | ···················· | ···················· | ···················· | ···················· |
| L | 2694 hCoV-19/New Zealand/20VR1830/2020\|EPI_ISL_579142\|2 | ···················· | ···················· | ···················· | ···················· | ···················· | ···················· |
| L | 2695 hCoV-19/New Zealand/20VR1835/2020\|EPI_ISL_579143\|2 | ···················· | ···················· | ···················· | ···················· | ···················· | ···················· |
| L | 2696 hCoV-19/New Zealand/20VR1840/2020\|EPI_ISL_579144\|2 | ···················· | ···················· | ···················· | ···················· | ···················· | ···················· |
| L | 2697 hCoV-19/New Zealand/20VR1842/2020\|EPI_ISL_579145\|2 | ···················· | ···················· | ···················· | ···················· | ···················· | ···················· |
| L | 2698 hCoV-19/New Zealand/20VR1844/2020\|EPI_ISL_579146\|2 | ···················· | ···················· | ···················· | ···················· | ···················· | ···················· |
| L | 2699 hCoV-19/New Zealand/20VR1846/2020\|EPI_ISL_579147\|2 | ···················· | ···················· | ···················· | ···················· | ···················· | ···················· |
| L | 2700 hCoV-19/New Zealand/20VR1848/2020\|EPI_ISL_579148\|2 | ···················· | ···················· | ···················· | ···················· | ···················· | ···················· |
| L | 2702 hCoV-19/New Zealand/20VR1851/2020\|EPI_ISL_579150\|2 | ···················· | ···················· | ···················· | ···················· | ···················· | ···················· |
| L | 2703 hCoV-19/New Zealand/20VR1852/2020\|EPI_ISL_579151\|2 | ···················· | ···················· | ···················· | ···················· | ···················· | ···················· |
| L | 2704 hCoV-19/New Zealand/20VR1853/2020\|EPI_ISL_579152\|2 | ···················· | ···················· | ···················· | ···················· | ···················· | ···················· |
| L | 2705 hCoV-19/New Zealand/20VR1859/2020\|EPI_ISL_579153\|2 | ···················· | ···················· | ···················· | ···················· | ···················· | ···················· |
| L | 2706 hCoV-19/New Zealand/20VR1861/2020\|EPI_ISL_579154\|2 | ···················· | ···················· | ···················· | ···················· | ···················· | ···················· |
| L | 2707 hCoV-19/New Zealand/20VR1868/2020\|EPI_ISL_579155\|2 | ···················· | ···················· | ···················· | ···················· | ···················· | ···················· |
| L | 2708 hCoV-19/New Zealand/20VR1888/2020\|EPI_ISL_579156\|2 | ···················· | ···················· | ···················· | ···················· | ···················· | ···················· |
| L | 2709 hCoV-19/New Zealand/20VR1889/2020\|EPI_ISL_579157\|2 | ···················· | ···················· | ···················· | ···················· | ···················· | ···················· |
| L | 2710 hCoV-19/New Zealand/20VR1895/2020\|EPI_ISL_579158\|2 | ···················· | ···················· | ···················· | ···················· | ···················· | ···················· |
| L | 2711 hCoV-19/New Zealand/20VR1901/2020\|EPI_ISL_579159\|2 | ···················· | ···················· | ···················· | ···················· | ···················· | ···················· |
| L | 2712 hCoV-19/England/ALDP-9E9FDA/2020\|EPI_ISL_580100\|20 | ···················· | ·A·················· | ···················· | ···················· | ···················· | ···A················ |
| L | 2713 hCoV-19/England/ALDP-9EA1E2/2020\|EPI_ISL_580101\|20 | ···················· | ···················· | ···················· | ···················· | ···················· | ···················· |
| L | 2714 hCoV-19/England/ALDP-9EAB02/2020\|EPI_ISL_580103\|20 | ···················· | ···················· | ···················· | ···················· | ···················· | ···················· |
| L | 2715 hCoV-19/England/ALDP-9E9B7C/2020\|EPI_ISL_580104\|20 | ···················· | ···················· | ···················· | ···················· | ···················· | ···················· |
| L | 2716 hCoV-19/England/ALDP-9E9D0D/2020\|EPI_ISL_580105\|20 | ···················· | ···················· | ···················· | ···················· | ···················· | ···················· |
| L | 2717 hCoV-19/England/ALDP-9E9E73/2020\|EPI_ISL_580106\|20 | ···················· | ···················· | ···················· | ···················· | ···················· | ···················· |
| L | 2718 hCoV-19/England/ALDP-9E9B30/2020\|EPI_ISL_580107\|20 | ···················· | ·A·················· | ···················· | ···················· | ···················· | ···A················ |
| L | 2719 hCoV-19/England/ALDP-9E9F70/2020\|EPI_ISL_580108\|20 | ···················· | ···················· | ···················· | ···················· | ···················· | ···················· |
| L | 2720 hCoV-19/England/ALDP-9E9DFE/2020\|EPI_ISL_580109\|20 | ···················· | ···················· | ···················· | ···················· | ···················· | ···················· |
| L | 2721 hCoV-19/England/ALDP-9EA6A7/2020\|EPI_ISL_580110\|20 | ···················· | ···················· | ···················· | ···················· | ···················· | ···················· |
| L | 2722 hCoV-19/England/ALDP-9EAFF7/2020\|EPI_ISL_580111\|20 | ···················· | ···················· | ···················· | ···················· | ···················· | ···················· |
| L | 2723 hCoV-19/England/ALDP-9E9DD0/2020\|EPI_ISL_580112\|20 | ···················· | ···················· | ···················· | ···················· | ···················· | ···················· |
| L | 2724 hCoV-19/England/ALDP-9EA14C/2020\|EPI_ISL_580113\|20 | ···················· | ···················· | ···················· | ···················· | ···················· | ···················· |
| L | 2725 hCoV-19/England/ALDP-9EA319/2020\|EPI_ISL_580115\|20 | ···················· | ···················· | ···················· | ···················· | ···················· | ···················· |
| L | 2726 hCoV-19/England/ALDP-9EAFAC/2020\|EPI_ISL_580116\|20 | ···················· | ···················· | ···················· | ···················· | ···················· | ···················· |
| L | 2727 hCoV-19/England/ALDP-9EA67A/2020\|EPI_ISL_580119\|20 | ···················· | ···················· | ···················· | ···················· | ···················· | ···················· |
| L | 2728 hCoV-19/England/ALDP-9E9C00/2020\|EPI_ISL_580126\|20 | ···················· | ···················· | ···················· | ···················· | ···················· | ···················· |
| L | 2729 hCoV-19/England/ALDP-9EAD48/2020\|EPI_ISL_580142\|20 | ···················· | ···················· | ···················· | ···················· | ······T············· | ···················· |
| L | 2730 hCoV-19/England/ALDP-9E9CE2/2020\|EPI_ISL_580143\|20 | ···················· | ···················· | ···················· | ···················· | ···················· | ···················· |
| L | 2731 hCoV-19/England/ALDP-9E9DA3/2020\|EPI_ISL_580145\|20 | ···················· | ······A············· | ···················· | ···················· | ····T··············· | ···················· |
| L | 2732 hCoV-19/England/ALDP-9EA461/2020\|EPI_ISL_580146\|20 | ···················· | ···················· | ···················· | ···················· | ···················· | ···················· |
| L | 2733 hCoV-19/England/ALDP-9EA64D/2020\|EPI_ISL_580147\|20 | ···················· | ···················· | ···················· | ···················· | ···················· | ···················· |
| L | 2734 hCoV-19/England/ALDP-9EA66B/2020\|EPI_ISL_580149\|20 | ···················· | ···················· | ···················· | ···················· | ······T············· | ···················· |
| L | 2735 hCoV-19/England/ALDP-9EA16A/2020\|EPI_ISL_580151\|20 | ···················· | ···················· | ···················· | ···················· | ···················· | ···················· |
| L | 2736 hCoV-19/England/ALDP-9EAD84/2020\|EPI_ISL_580152\|20 | ···················· | ···················· | ···················· | ···················· | ···················· | ···················· |
| L | 2737 hCoV-19/England/ALDP-9E78D1/2020\|EPI_ISL_580153\|20 | ···················· | ···················· | ···················· | ···················· | ···················· | ···················· |
| L | 2738 hCoV-19/England/ALDP-9EA6B6/2020\|EPI_ISL_580154\|20 | ···················· | ···················· | ···················· | ···················· | ···················· | ···················· |
| L | 2739 hCoV-19/England/ALDP-9EAB3F/2020\|EPI_ISL_580155\|20 | ···················· | ···················· | ···················· | ···················· | ···················· | ···················· |
| L | 2740 hCoV-19/England/ALDP-9EA55F/2020\|EPI_ISL_580156\|20 | ···················· | ·A·················· | ···················· | ···················· | ···················· | ···················· |
| L | 2741 hCoV-19/England/ALDP-9E9AE8/2020\|EPI_ISL_580157\|20 | ···················· | ···················· | ···················· | ···················· | ···················· | ···················· |
| L | 2742 hCoV-19/England/ALDP-9EAABA/2020\|EPI_ISL_580158\|20 | ···················· | ···················· | ···················· | ···················· | ···················· | ···················· |
| L | 2743 hCoV-19/England/ALDP-9EA6D4/2020\|EPI_ISL_580159\|20 | ···················· | ···················· | ···················· | ···················· | ···················· | ···················· |
| L | 2744 hCoV-19/England/ALDP-9E9B8B/2020\|EPI_ISL_580160\|20 | ···················· | ···················· | ···················· | ···················· | ···················· | ···················· |
| L | 2745 hCoV-19/England/ALDP-9EAF7F/2020\|EPI_ISL_580161\|20 | ···················· | ···················· | ···················· | ···················· | ···················· | ···················· |
| L | 2746 hCoV-19/England/ALDP-9E7288/2020\|EPI_ISL_580291\|20 | ···················· | ···················· | ···················· | ···················· | ···················· | ···················· |
| L | 2747 hCoV-19/England/ALDP-9E70CA/2020\|EPI_ISL_580296\|20 | ···················· | ···················· | ···················· | ···················· | ···················· | ···················· |
| L | 2748 hCoV-19/England/ALDP-9E70E8/2020\|EPI_ISL_580297\|20 | ···················· | ···················· | ···················· | ···················· | ···················· | ···················· |
| L | 2749 hCoV-19/England/ALDP-9E684B/2020\|EPI_ISL_580298\|20 | ···················· | ···················· | ···················· | ···················· | ···················· | ···················· |
| L | 2750 hCoV-19/England/CAMC-9EC283/2020\|EPI_ISL_580299\|20 | ···················· | ···················· | ···················· | ···················· | ···················· | ···················· |
| L | 2751 hCoV-19/England/QEUH-962461/2020\|EPI_ISL_581410\|20 | ···················· | ···················· | ···················· | ···················· | ···················· | ···················· |

| | | | | | | | | |
|---|---|---|---|---|---|---|---|---|
| L | 2752 | hCoV-19/England/ALDP-95574A/2020\|EPI_ISL_581411\|20 | .................... | .................... | .................... | .................... | .................... | .................... |
| L | 2753 | hCoV-19/England/ALDP-95074F/2020\|EPI_ISL_581412\|20 | .................... | .................... | .................... | .................... | .................... | .................... |
| L | 2754 | hCoV-19/England/ALDP-94FF87/2020\|EPI_ISL_581413\|20 | .................... | .................... | .................... | .................... | .................... | .................... |
| L | 2755 | hCoV-19/England/ALDP-95016F/2020\|EPI_ISL_581414\|20 | .................... | .................... | .................... | .................... | .................... | .................... |
| L | 2756 | hCoV-19/England/ALDP-95C2E8/2020\|EPI_ISL_581416\|20 | .................... | .................... | .................... | .................... | .................... | .................... |
| L | 2757 | hCoV-19/England/ALDP-95AD61/2020\|EPI_ISL_581417\|20 | .................... | .................... | .................... | .................... | .................... | .................... |
| L | 2758 | hCoV-19/England/QEUH-960A9E/2020\|EPI_ISL_581418\|20 | .................... | .................... | .................... | .................... | .................... | .................... |
| L | 2759 | hCoV-19/England/QEUH-960B13/2020\|EPI_ISL_581419\|20 | .................... | .................... | .................... | .................... | .................... | .................... |
| L | 2760 | hCoV-19/England/QEUH-960A52/2020\|EPI_ISL_581420\|20 | .................... | .................... | .................... | .................... | .................... | .................... |
| L | 2761 | hCoV-19/England/MILK-95DB82/2020\|EPI_ISL_581421\|20 | .................... | .................... | .................... | .................... | .................... | .................... |
| L | 2762 | hCoV-19/England/MILK-95DBCE/2020\|EPI_ISL_581422\|20 | .................... | .................... | .................... | .................... | .................... | .................... |
| L | 2763 | hCoV-19/England/ALDP-8D1B96/2020\|EPI_ISL_581426\|20 | .................... | .................... | .................... | .................... | .................... | .................... |
| L | 2764 | hCoV-19/England/ALDP-6FEA6B/2020\|EPI_ISL_581427\|20 | .................... | .................... | .................... | .................... | .................... | .................... |
| L | 2765 | hCoV-19/England/MILK-863258/2020\|EPI_ISL_581428\|20 | .................... | .................... | .................... | .................... | .................... | .................... |
| L | 2766 | hCoV-19/England/ALDP-6FA9BD/2020\|EPI_ISL_581429\|20 | .................... | .................... | .................... | .................... | .................... | .................... |
| L | 2767 | hCoV-19/England/ALDP-705796/2020\|EPI_ISL_581430\|20 | .................... | .................... | .................... | .................... | .................... | .................... |
| L | 2768 | hCoV-19/England/ALDP-702641/2020\|EPI_ISL_581431\|20 | .................... | .................... | .................... | .................... | .................... | .................... |
| L | 2769 | hCoV-19/England/ALDP-71FB7F/2020\|EPI_ISL_581432\|20 | .................... | .............A...... | .................... | .................... | .................... | .................... |
| L | 2770 | hCoV-19/England/MILK-60B70A/2020\|EPI_ISL_581434\|20 | .................... | .................... | .................... | .................... | .................... | .................... |
| L | 2771 | hCoV-19/England/ALDP-643DF5/2020\|EPI_ISL_581435\|20 | .................... | .................... | .................... | .................... | .................... | .................... |
| L | 2772 | hCoV-19/England/ALDP-59A15E/2020\|EPI_ISL_581436\|20 | .................... | .................... | .................... | .................... | .................... | .................... |
| L | 2773 | hCoV-19/England/QEUH-5ED462/2020\|EPI_ISL_581437\|20 | .................... | .................... | .................... | .................... | .................... | .................... |
| L | 2774 | hCoV-19/England/MILK-574DFC/2020\|EPI_ISL_581438\|20 | .................... | .................... | .................... | .................... | .................... | .................... |
| L | 2775 | hCoV-19/England/QEUH-5ED5D8/2020\|EPI_ISL_581439\|20 | .................... | .................... | .................... | .................... | .................... | .................... |
| L | 2776 | hCoV-19/England/QEUH-5ED5F6/2020\|EPI_ISL_581441\|20 | .................... | .................... | .................... | .................... | .................... | .................... |
| L | 2777 | hCoV-19/England/ALDP-5D00E6/2020\|EPI_ISL_581442\|20 | .................... | .................... | .................... | .................... | .................... | .................... |
| L | 2778 | hCoV-19/England/QEUH-5DB537/2020\|EPI_ISL_581443\|20 | .................... | .................... | .................... | .................... | .................... | .................... |
| L | 2779 | hCoV-19/England/ALDP-521AF7/2020\|EPI_ISL_581444\|20 | .................... | .................... | .................... | .................... | .................... | .................... |
| L | 2780 | hCoV-19/England/ALDP-4F393E/2020\|EPI_ISL_581446\|20 | .................... | .................... | .................... | .................... | .................... | .................... |
| L | 2781 | hCoV-19/England/MILK-4BC157/2020\|EPI_ISL_581447\|20 | .................... | .................... | .................... | .................... | .................... | .................... |
| L | 2782 | hCoV-19/England/ALDP-4F3619/2020\|EPI_ISL_581448\|20 | .................... | .................... | .................... | .................... | .................... | .................... |
| L | 2783 | hCoV-19/India/WB-IICB-017/2020\|EPI_ISL_581449\|2020 | .................... | .................... | .................... | .................... | .................... | .................... |
| L | 2784 | hCoV-19/India/WB-IICB-018/2020\|EPI_ISL_581450\|2020 | .................... | .................... | .................... | .................... | .................... | .................... |
| L | 2785 | hCoV-19/Germany/BW-UKT-034/2020\|EPI_ISL_581478\|202 | .................... | .................... | .................... | .................... | .................... | .................... |
| L | 2786 | hCoV-19/Germany/BW-UKT-036/2020\|EPI_ISL_581479\|202 | .................... | .................... | .................... | .................... | .................... | .................... |
| L | 2787 | hCoV-19/Germany/BW-UKT-039/2020\|EPI_ISL_581480\|202 | .................... | .................... | .................... | .................... | .................... | .................... |
| L | 2788 | hCoV-19/Germany/BW-UKT-044/2020\|EPI_ISL_581481\|202 | .................... | .................... | .................... | .................... | .................... | .................... |
| L | 2789 | hCoV-19/Germany/BW-UKT-045/2020\|EPI_ISL_581482\|202 | .................... | .................... | .................... | .................... | .................... | .................... |
| L | 2790 | hCoV-19/Germany/BW-UKT-048/2020\|EPI_ISL_581483\|202 | .................... | .................... | .................... | .................... | .................... | .................... |
| L | 2791 | hCoV-19/Germany/BW-UKT-051/2020\|EPI_ISL_581484\|202 | .................... | .................... | .................... | .................... | .................... | .................... |
| L | 2792 | hCoV-19/Germany/BW-UKT-062/2020\|EPI_ISL_581485\|202 | .................... | .................... | .................... | .................... | .................... | .................... |
| L | 2793 | hCoV-19/Congo/UKT-001/2020\|EPI_ISL_581486\|2020-04 | .................... | .................... | .................... | .................... | .................... | .................... |
| L | 2794 | hCoV-19/Congo/UKT-002/2020\|EPI_ISL_581487\|2020-07- | .................... | .................... | .................... | .................... | .................... | .................... |
| L | 2795 | hCoV-19/Congo/UKT-004/2020\|EPI_ISL_581488\|2020-05 | .................... | .................... | .................... | .................... | .................... | .................... |
| L | 2796 | hCoV-19/Congo/UKT-005/2020\|EPI_ISL_581489\|2020-06- | .................... | .................... | .................... | .................... | .................... | .................... |
| L | 2797 | hCoV-19/Congo/UKT-009/2020\|EPI_ISL_581490\|2020-06- | .................... | .................... | .................... | .................... | .................... | .................... |
| L | 2798 | hCoV-19/Congo/UKT-013/2020\|EPI_ISL_581491\|2020-06- | .................... | .................... | .................... | .................... | .................... | .................... |
| L | 2799 | hCoV-19/Congo/UKT-014/2020\|EPI_ISL_581492\|2020-07- | .................... | .................... | .................... | .................... | .................... | .................... |
| L | 2800 | hCoV-19/Congo/UKT-015/2020\|EPI_ISL_581493\|2020-06- | .................... | .................... | .................... | .................... | .................... | .................... |
| L | 2801 | hCoV-19/India/WB-IICB-020/2020\|EPI_ISL_581494\|2020 | .................... | .................... | .................... | .................... | .................... | .................... |
| L | 2802 | hCoV-19/Italy/SIC-IZSBM169-66/2020\|EPI_ISL_582767\| | .................... | .................... | .................... | .................... | .................... | .................... |
| L | 2803 | hCoV-19/Italy/SIC-IZSBM169-55/2020\|EPI_ISL_582769\| | .................... | .................... | .................... | .................... | .................... | .................... |
| L | 2804 | hCoV-19/Sweden/20-08985/2020\|EPI_ISL_582771\|2020-0 | .................... | .................... | .................... | .................... | .................... | .................... |
| L | 2805 | hCoV-19/Sweden/20-08990/2020\|EPI_ISL_582772\|2020-0 | .................... | .................... | .................... | .................... | .................... | .................... |
| L | 2806 | hCoV-19/Sweden/20-08991/2020\|EPI_ISL_582773\|2020-0 | .................... | .................... | .................... | .................... | .................... | .................... |
| L | 2807 | hCoV-19/Sweden/20-08992/2020\|EPI_ISL_582774\|2020-0 | .................... | .................... | .................... | .................... | .................... | .................... |
| L | 2808 | hCoV-19/Sweden/20-08993/2020\|EPI_ISL_582775\|2020-0 | .................... | .................... | .................... | .................... | .................... | .................... |
| L | 2809 | hCoV-19/Sweden/20-21497/2020\|EPI_ISL_582776\|2020-0 | .................... | .................... | .................... | .................... | .................... | .................... |
| L | 2810 | hCoV-19/Sweden/20-21498/2020\|EPI_ISL_582777\|2020-0 | .................... | .................... | .................... | .................... | .................... | .................... |
| L | 2811 | hCoV-19/Sweden/20-21500/2020\|EPI_ISL_582778\|2020-0 | .................... | .................... | .................... | .................... | .................... | .................... |
| L | 2812 | hCoV-19/Sweden/20-21501/2020\|EPI_ISL_582779\|2020-0 | .................... | .................... | .................... | .................... | .................... | .................... |
| L | 2813 | hCoV-19/Sweden/20-21506/2020\|EPI_ISL_582780\|2020-0 | .................... | .................... | .................... | .................... | .................... | .................... |
| L | 2814 | hCoV-19/Sweden/20-21507/2020\|EPI_ISL_582781\|2020-0 | .................... | .................... | .................... | .................... | .................... | .................... |
| L | 2815 | hCoV-19/Sweden/20-22114/2020\|EPI_ISL_582788\|2020-0 | .................... | .................... | .................... | .................... | .................... | .................... |
| L | 2816 | hCoV-19/Sweden/20-22115/2020\|EPI_ISL_582789\|2020-0 | .................... | .................... | .................... | .................... | .................... | .................... |
| L | 2817 | hCoV-19/England/ALDP-9B6352/2020\|EPI_ISL_573000\|20 | .................... | .................... | .................... | .................... | .................... | .................... |
| L | 2818 | hCoV-19/England/ALDP-9B639E/2020\|EPI_ISL_573001\|20 | .................... | .................... | .................... | .................... | .................... | .................... |
| L | 2819 | hCoV-19/England/ALDP-9B63AD/2020\|EPI_ISL_573002\|20 | .................... | .................... | .................... | .................... | .................... | .................... |
| L | 2820 | hCoV-19/England/ALDP-9B63E9/2020\|EPI_ISL_573003\|20 | .................... | .................... | .................... | .................... | .................... | .................... |
| L | 2821 | hCoV-19/England/ALDP-9B6422/2020\|EPI_ISL_573004\|20 | .................... | .................... | .................... | .................... | .................... | .................... |
| L | 2822 | hCoV-19/England/ALDP-9B6431/2020\|EPI_ISL_573005\|20 | .................... | .................... | .................... | .................... | .................... | .................... |
| L | 2823 | hCoV-19/England/ALDP-9B6440/2020\|EPI_ISL_573006\|20 | .................... | .................... | .................... | .................... | .................... | .................... |
| L | 2824 | hCoV-19/England/ALDP-9B645F/2020\|EPI_ISL_573007\|20 | .................... | .................... | .................... | .................... | .................... | .................... |
| L | 2825 | hCoV-19/England/ALDP-9B646E/2020\|EPI_ISL_573008\|20 | .................... | .................... | .................... | .................... | .................... | .................... |
| L | 2826 | hCoV-19/England/ALDP-9B647D/2020\|EPI_ISL_573009\|20 | .................... | .................... | .................... | .................... | .................... | .................... |
| L | 2827 | hCoV-19/England/ALDP-9B64B9/2020\|EPI_ISL_573010\|20 | .................... | .................... | .................... | .................... | .................... | .................... |
| L | 2828 | hCoV-19/England/ALDP-9B64D7/2020\|EPI_ISL_573011\|20 | .................... | .................... | .................... | .................... | .................... | .................... |
| L | 2829 | hCoV-19/England/ALDP-9B64E6/2020\|EPI_ISL_573012\|20 | .................... | .................... | .................... | .................... | .................... | .................... |
| L | 2830 | hCoV-19/England/ALDP-9B64F5/2020\|EPI_ISL_573013\|20 | .................... | .................... | .................... | .................... | .................... | .................... |
| L | 2831 | hCoV-19/England/ALDP-9B653E/2020\|EPI_ISL_573014\|20 | .................... | .................... | .................... | .................... | .................... | .................... |

```
L 2832 hCoV-19/England/ALDP-9B655C/2020|EPI_ISL_573015|20 .................... .A.................. ..................... .................... .................... ....................
L 2833 hCoV-19/England/ALDP-9B656B/2020|EPI_ISL_573016|20 .................... .................... ..................... .................... .................... ....................
L 2834 hCoV-19/England/ALDP-9B65A7/2020|EPI_ISL_573017|20 .................... .................... ..................... .................... .................... ....................
L 2835 hCoV-19/England/ALDP-9B65D4/2020|EPI_ISL_573018|20 .................... .A.................. ..................... .................... .................... ....................
L 2836 hCoV-19/England/ALDP-9B65E3/2020|EPI_ISL_573019|20 .................... .................... ..................... .................... .................... ....................
L 2837 hCoV-19/England/ALDP-9B661D/2020|EPI_ISL_573020|20 .................... .................... ..................... .................... .................... ....................
L 2838 hCoV-19/England/ALDP-9B663B/2020|EPI_ISL_573021|20 .................... .................... ..................... .................... .................... ....................
L 2839 hCoV-19/England/ALDP-9B664A/2020|EPI_ISL_573022|20 .................... .................... ..................... .................... .................... ....................
L 2840 hCoV-19/England/ALDP-9B6659/2020|EPI_ISL_573023|20 .................... .................... ..................... .................... .................... ....................
L 2841 hCoV-19/England/ALDP-9B6668/2020|EPI_ISL_573024|20 .................... .................... ..................... .................... .................... ....................
L 2842 hCoV-19/England/ALDP-9B6677/2020|EPI_ISL_573025|20 .................... .................... ..................... .................... .................... ....................
L 2843 hCoV-19/England/ALDP-9B6686/2020|EPI_ISL_573026|20 .................... .................... ..................... .................... .................... ....................
L 2844 hCoV-19/England/ALDP-9B6695/2020|EPI_ISL_573027|20 .................... .................... ..................... .................... .................... ....................
L 2845 hCoV-19/England/ALDP-9B66D1/2020|EPI_ISL_573028|20 .................... .................... ..................... .................... .................... ....................
L 2846 hCoV-19/England/ALDP-9B66E0/2020|EPI_ISL_573029|20 .................... .................... ..................... .................... .................... ....................
L 2847 hCoV-19/England/ALDP-9B66FF/2020|EPI_ISL_573030|20 .................... .................... ..................... .................... .................... ....................
L 2848 hCoV-19/England/ALDP-9B6729/2020|EPI_ISL_573031|20 .................... .................... ..................... .................... .................... ....................
L 2849 hCoV-19/England/ALDP-9B6738/2020|EPI_ISL_573032|20 .................... .................... ..................... .................... .................... ....................
L 2850 hCoV-19/England/ALDP-9B6756/2020|EPI_ISL_573033|20 .................... .................... ..................... .................... .................... ....................
L 2851 hCoV-19/England/ALDP-9B6765/2020|EPI_ISL_573034|20 .................... .................... ..................... .................... .................... ....................
L 2852 hCoV-19/England/ALDP-9B6783/2020|EPI_ISL_573035|20 .................... .................... ..................... .................... .................... ....................
L 2853 hCoV-19/England/ALDP-9B6792/2020|EPI_ISL_573036|20 .................... .................... ..................... .................... .................... ....................
L 2854 hCoV-19/England/ALDP-9B6808/2020|EPI_ISL_573037|20 .................... .A.................. ..................... .................... .................... ...A................
L 2855 hCoV-19/England/ALDP-9BC9A5/2020|EPI_ISL_573038|20 .................... .................... ..................... .................... .................... ....................
L 2856 hCoV-19/England/ALDP-9BC9B4/2020|EPI_ISL_573039|20 .................... .................... ..................... .................... .................... ....................
L 2857 hCoV-19/England/ALDP-9BC9E1/2020|EPI_ISL_573040|20 .................... .................... ..................... .................... .................... ....................
L 2858 hCoV-19/England/ALDP-9BCA2A/2020|EPI_ISL_573041|20 .................... .................... ..................... .................... .................... ....................
L 2859 hCoV-19/England/ALDP-9BCA48/2020|EPI_ISL_573042|20 .................... .................... ..................... .................... .................... ....................
L 2860 hCoV-19/England/ALDP-9BCA57/2020|EPI_ISL_573043|20 .................... .................... ..................... .................... .................... ....................
L 2861 hCoV-19/England/ALDP-9BCA66/2020|EPI_ISL_573044|20 .................... .A.................. ..................... .................... .................... ...A................
L 2862 hCoV-19/England/ALDP-9BCA75/2020|EPI_ISL_573045|20 .................... .A.................. ..................... .................... .................... ...A................
L 2863 hCoV-19/England/ALDP-9BCA93/2020|EPI_ISL_573046|20 .................... .................... ...........T......... .................... .................... ....................
L 2864 hCoV-19/England/ALDP-9BCAA2/2020|EPI_ISL_573047|20 .................... .................... ..................... .................... .................... ....................
L 2865 hCoV-19/England/ALDP-9BCAB1/2020|EPI_ISL_573048|20 .................... .................... ..................... .................... .................... ....................
L 2866 hCoV-19/England/ALDP-9BCAC0/2020|EPI_ISL_573049|20 .................... .................... ..................... .................... .................... ....................
L 2867 hCoV-19/England/ALDP-9BCAFD/2020|EPI_ISL_573050|20 .................... .................... ..................... .................... .................... ....................
L 2868 hCoV-19/England/ALDP-9BCB18/2020|EPI_ISL_573051|20 .................... .................... ..................... .................... .................... ....................
L 2869 hCoV-19/England/ALDP-9BCB27/2020|EPI_ISL_573052|20 .................... .................... ..................... .................... .................... ....................
L 2870 hCoV-19/England/ALDP-9BCB36/2020|EPI_ISL_573053|20 .................... .A.................. ..................... .................... .................... ...A................
L 2871 hCoV-19/England/ALDP-9BCB45/2020|EPI_ISL_573054|20 .................... .................... ..................... .................... .................... ....................
L 2872 hCoV-19/England/ALDP-9BCB54/2020|EPI_ISL_573055|20 .................... .A.................. ..................... .................... .................... ...A................
L 2873 hCoV-19/England/ALDP-9BCB72/2020|EPI_ISL_573056|20 .................... .................... ..................... .................... .................... ....................
L 2874 hCoV-19/England/ALDP-9BCB81/2020|EPI_ISL_573057|20 .................... .................... ..................... .................... .................... ....................
L 2875 hCoV-19/England/ALDP-9BCBDC/2020|EPI_ISL_573061|20 .................... .................... ..................... .................... .................... ....................
L 2876 hCoV-19/England/ALDP-9BCBFA/2020|EPI_ISL_573063|20 .................... .................... ..................... .................... .................... ....................
L 2877 hCoV-19/England/ALDP-9BCC06/2020|EPI_ISL_573064|20 .................... .................... ..................... .................... .................... ....................
L 2878 hCoV-19/England/ALDP-9BCCBB/2020|EPI_ISL_573069|20 .................... .................... ..................... .................... .................... ....................
L 2879 hCoV-19/England/ALDP-9BCCD9/2020|EPI_ISL_573070|20 .................... .A.................. ..................... .................... .................... ...A................
L 2880 hCoV-19/England/ALDP-9BCCE8/2020|EPI_ISL_573071|20 .................... .................... ..................... .................... .................... ....................
L 2881 hCoV-19/England/ALDP-9BCD30/2020|EPI_ISL_573072|20 .................... .................... ..................... .................... .................... ....................
L 2882 hCoV-19/England/ALDP-9BCD5E/2020|EPI_ISL_573073|20 .................... .................... ..................... .................... .................... ....................
L 2883 hCoV-19/England/ALDP-9BCD6D/2020|EPI_ISL_573074|20 .................... .................... ..................... .................... .................... ....................
L 2884 hCoV-19/England/ALDP-9BCD9A/2020|EPI_ISL_573075|20 .................... .A.................. ..................... .................... .................... ...A................
L 2885 hCoV-19/England/ALDP-9BCDF4/2020|EPI_ISL_573076|20 .................... .................... ..................... .................... .................... ....................
L 2886 hCoV-19/England/ALDP-9BCE00/2020|EPI_ISL_573077|20 .................... .A.................. ..................... .................... .................... ...A................
L 2887 hCoV-19/England/ALDP-9BCE2E/2020|EPI_ISL_573078|20 .................... .................... ..................... .................... .................... ....................
L 2888 hCoV-19/England/ALDP-9BCE4C/2020|EPI_ISL_573079|20 .................... .................... ..................... .................... .................... ....................
L 2889 hCoV-19/USA/NM-DOH-2020315962/2020|EPI_ISL_574291| .................... ....A............... ..................... .................... .................... ....................
L 2890 hCoV-19/USA/LA-EVTL160/2020|EPI_ISL_574295|2020-05 .................... .................... ..................... .................... .................... ....................
L 2891 hCoV-19/USA/LA-EVTL161/2020|EPI_ISL_574296|2020-05 .................... .................... ..................... .................... .................... ....................
L 2892 hCoV-19/USA/LA-EVTL162/2020|EPI_ISL_574297|2020-05 .................... .................... ..................... .................... .................... ....................
L 2893 hCoV-19/USA/LA-EVTL163/2020|EPI_ISL_574298|2020-05 .................... .................... ..................... .................... .................... ....................
L 2894 hCoV-19/USA/LA-EVTL164/2020|EPI_ISL_574299|2020-05 .................... .................... ..................... .................... .................... ....................
L 2895 hCoV-19/USA/LA-EVTL189/2020|EPI_ISL_574323|2020-05 .................... .................... ..................... .................... .................... ....................
L 2896 hCoV-19/USA/LA-EVTL190/2020|EPI_ISL_574324|2020-05 .................... .................... ..................... .................... .................... ....................
L 2897 hCoV-19/USA/LA-EVTL191/2020|EPI_ISL_574325|2020-05 .................... .................... ..................... .................... .................... ....................
L 2898 hCoV-19/USA/LA-EVTL192/2020|EPI_ISL_574326|2020-07 .................... .................... ..................... .................... .................... ....................
L 2899 hCoV-19/USA/LA-EVTL193/2020|EPI_ISL_574327|2020-07 .................... .................... ..................... .................... .................... ....................
L 2900 hCoV-19/USA/LA-EVTL194/2020|EPI_ISL_574328|2020-08 .................... .A.................. ..................... .................... .................... ....................
L 2901 hCoV-19/USA/LA-EVTL195/2020|EPI_ISL_574329|2020-08 .................... .................... ..................... .................... .................... ....................
L 2902 hCoV-19/USA/LA-EVTL196/2020|EPI_ISL_574330|2020-08 .................... .................... ..................... .................... .................... ....................
L 2903 hCoV-19/USA/LA-EVTL197/2020|EPI_ISL_574331|2020-08 .................... .................... ..................... .................... .................... ....................
L 2904 hCoV-19/USA/LA-EVTL198/2020|EPI_ISL_574332|2020-08 .................... .................... ..................... .................... .................... ....................
L 2905 hCoV-19/USA/LA-EVTL199/2020|EPI_ISL_574333|2020-08 .................... .................... ..................... .................... .................... ....................
L 2906 hCoV-19/USA/LA-EVTL200/2020|EPI_ISL_574334|2020-08 .................... .................... ..................... .................... .................... ....................
L 2907 hCoV-19/USA/LA-EVTL201/2020|EPI_ISL_574335|2020-08 .................... .................... ..................... .................... .................... ....................
L 2908 hCoV-19/USA/LA-EVTL202/2020|EPI_ISL_574336|2020-08 .................... .................... ..................... .................... .................... ....................
L 2909 hCoV-19/USA/LA-EVTL203/2020|EPI_ISL_574337|2020-08 .................... .................... ..................... .................... .................... ....................
L 2910 hCoV-19/USA/LA-EVTL204/2020|EPI_ISL_574338|2020-08 .................... .................... ..................... .................... .................... ....................
L 2911 hCoV-19/USA/LA-EVTL205/2020|EPI_ISL_574339|2020-08 .................... .................... ..................... .................... .................... ....................
```

| | | | | | | | | | |
|---|---|---|---|---|---|---|---|---|---|
| L | 2912 | hCoV-19/USA/LA-EVTL206/2020\|EPI_ISL_574340\|2020-08 | ···················· | ···················· | ···················· | ···················· | ···················· | ···················· | |
| L | 2913 | hCoV-19/USA/LA-EVTL207/2020\|EPI_ISL_574341\|2020-08 | ···················· | ···················· | ···················· | ···················· | ···················· | ···················· | |
| L | 2914 | hCoV-19/USA/LA-EVTL208/2020\|EPI_ISL_574342\|2020-08 | ···················· | ···················· | ···················· | ···················· | ···················· | ···················· | |
| L | 2915 | hCoV-19/USA/LA-EVTL209/2020\|EPI_ISL_574343\|2020-08 | ···················· | ···················· | ···················· | ···················· | ···················· | ···················· | |
| L | 2916 | hCoV-19/USA/LA-EVTL210/2020\|EPI_ISL_574344\|2020-08 | ···················· | ···················· | ···················· | ···················· | ···················· | ···················· | |
| L | 2917 | hCoV-19/USA/LA-EVTL211/2020\|EPI_ISL_574345\|2020-08 | ···················· | ···················· | ···················· | ···················· | ···················· | ···················· | |
| L | 2918 | hCoV-19/USA/LA-EVTL212/2020\|EPI_ISL_574346\|2020-08 | ···················· | ···················· | ···················· | ···················· | ···················· | ···················· | |
| L | 2919 | hCoV-19/USA/LA-EVTL213/2020\|EPI_ISL_574347\|2020-08 | ···················· | ···················· | ···················· | ···················· | ···················· | ···················· | |
| L | 2920 | hCoV-19/USA/LA-EVTL214/2020\|EPI_ISL_574348\|2020-08 | ···················· | ···················· | ···················· | ···················· | ···················· | ···················· | |
| L | 2921 | hCoV-19/USA/LA-EVTL215/2020\|EPI_ISL_574349\|2020-08 | ···················· | ···················· | ···················· | ···················· | ···················· | ···················· | |
| L | 2922 | hCoV-19/USA/LA-EVTL216/2020\|EPI_ISL_574350\|2020-08 | ···················· | ···················· | ···················· | ···················· | ···················· | ···················· | |
| L | 2923 | hCoV-19/USA/LA-EVTL217/2020\|EPI_ISL_574351\|2020-08 | ···················· | ···················· | ···················· | ···················· | ···················· | ···················· | |
| L | 2924 | hCoV-19/USA/LA-EVTL218/2020\|EPI_ISL_574352\|2020-08 | ···················· | ···················· | ···················· | ···················· | ···················· | ···················· | |
| L | 2925 | hCoV-19/USA/LA-EVTL219/2020\|EPI_ISL_574353\|2020-08 | ···················· | ···················· | ···················· | ···················· | ···················· | ···················· | |
| L | 2926 | hCoV-19/USA/LA-EVTL220/2020\|EPI_ISL_574354\|2020-08 | ···················· | ···················· | ···················· | ···················· | ···················· | ···················· | |
| L | 2927 | hCoV-19/USA/LA-EVTL221/2020\|EPI_ISL_574355\|2020-08 | ···················· | ···················· | ···················· | ···················· | ···················· | ···················· | |
| L | 2928 | hCoV-19/USA/LA-EVTL222/2020\|EPI_ISL_574356\|2020-08 | ···················· | ···················· | ···················· | ···················· | ···················· | ···················· | |
| L | 2929 | hCoV-19/USA/LA-EVTL223/2020\|EPI_ISL_574357\|2020-08 | ···················· | ···················· | ···················· | ···················· | ···················· | ···················· | |
| L | 2930 | hCoV-19/USA/LA-EVTL224/2020\|EPI_ISL_574358\|2020-08 | ···················· | ···················· | ···················· | ···················· | ···················· | ···················· | |
| L | 2931 | hCoV-19/USA/LA-EVTL225/2020\|EPI_ISL_574359\|2020-08 | ···················· | ···················· | ···················· | ···················· | ···················· | ···················· | |
| L | 2932 | hCoV-19/USA/LA-EVTL226/2020\|EPI_ISL_574360\|2020-08 | ···················· | ···················· | ···················· | ···················· | ···················· | ···················· | |
| L | 2933 | hCoV-19/USA/LA-EVTL227/2020\|EPI_ISL_574361\|2020-08 | ···················· | ···················· | ···················· | ···················· | ···················· | ···················· | |
| L | 2934 | hCoV-19/USA/LA-EVTL228/2020\|EPI_ISL_574362\|2020-08 | ···················· | ···················· | ···················· | ···················· | ···················· | ···················· | |
| L | 2935 | hCoV-19/USA/LA-EVTL229/2020\|EPI_ISL_574363\|2020-08 | ···················· | ···················· | ···················· | ···················· | ···················· | ···················· | |
| L | 2936 | hCoV-19/USA/LA-EVTL230/2020\|EPI_ISL_574364\|2020-08 | ···················· | ···················· | ···················· | ···················· | ···················· | ···················· | |
| L | 2937 | hCoV-19/USA/LA-EVTL231/2020\|EPI_ISL_574365\|2020-08 | ···················· | ···················· | ···················· | ···················· | ···················· | ···················· | |
| L | 2938 | hCoV-19/USA/LA-EVTL232/2020\|EPI_ISL_574366\|2020-08 | ···················· | ···················· | ···················· | ···················· | ···················· | ···················· | |
| L | 2939 | hCoV-19/USA/LA-EVTL233/2020\|EPI_ISL_574367\|2020-08 | ···················· | ···················· | ···················· | ···················· | ···················· | ···················· | |
| L | 2940 | hCoV-19/USA/LA-EVTL234/2020\|EPI_ISL_574368\|2020-08 | ···················· | ···················· | ···················· | ···················· | ···················· | ···················· | |
| L | 2941 | hCoV-19/USA/LA-EVTL235/2020\|EPI_ISL_574369\|2020-08 | ···················· | ···················· | ···················· | ···················· | ···················· | ···················· | |
| L | 2942 | hCoV-19/USA/LA-EVTL236/2020\|EPI_ISL_574370\|2020-08 | ···················· | ···················· | ···················· | ···················· | ···················· | ···················· | |
| L | 2943 | hCoV-19/USA/LA-EVTL237/2020\|EPI_ISL_574371\|2020-08 | ···················· | ···················· | ···················· | ···················· | ···················· | ···················· | |
| L | 2944 | hCoV-19/USA/LA-EVTL238/2020\|EPI_ISL_574372\|2020-08 | ···················· | ···················· | ···················· | ···················· | ···················· | ···················· | |
| L | 2945 | hCoV-19/USA/LA-EVTL239/2020\|EPI_ISL_574373\|2020-08 | ···················· | ···················· | ···················· | ···················· | ···················· | ···················· | |
| L | 2946 | hCoV-19/USA/LA-EVTL240/2020\|EPI_ISL_574374\|2020-08 | ···················· | ···················· | ···················· | ···················· | ···················· | ···················· | |
| L | 2947 | hCoV-19/USA/LA-EVTL241/2020\|EPI_ISL_574375\|2020-08 | ···················· | ···················· | ···················· | ···················· | ···················· | ···················· | |
| L | 2948 | hCoV-19/USA/LA-EVTL242/2020\|EPI_ISL_574376\|2020-08 | ···················· | ···················· | ···················· | ···················· | ···················· | ···················· | |
| L | 2949 | hCoV-19/USA/LA-EVTL243/2020\|EPI_ISL_574377\|2020-08 | ···················· | ···················· | ···················· | ···················· | ···················· | ···················· | |
| L | 2950 | hCoV-19/USA/LA-EVTL244/2020\|EPI_ISL_574378\|2020-08 | ···················· | ···················· | ···················· | ···················· | ···················· | ···················· | |
| L | 2951 | hCoV-19/USA/LA-EVTL245/2020\|EPI_ISL_574379\|2020-08 | ···················· | ···················· | ···················· | ···················· | ···················· | ···················· | |
| L | 2952 | hCoV-19/USA/LA-EVTL246/2020\|EPI_ISL_574380\|2020-08 | ···················· | ···················· | ···················· | ···················· | ···················· | ···················· | |
| L | 2953 | hCoV-19/USA/LA-EVTL247/2020\|EPI_ISL_574381\|2020-08 | ···················· | ···················· | ···················· | ···················· | ···················· | ···················· | |
| L | 2954 | hCoV-19/USA/LA-EVTL248/2020\|EPI_ISL_574382\|2020-08 | ···················· | ···················· | ···················· | ···················· | ···················· | ···················· | |
| L | 2955 | hCoV-19/USA/LA-EVTL249/2020\|EPI_ISL_574383\|2020-08 | ···················· | ···················· | ···················· | ···················· | ···················· | ···················· | |
| L | 2956 | hCoV-19/USA/LA-EVTL250/2020\|EPI_ISL_574384\|2020-08 | ···················· | ···················· | ···················· | ···················· | ···················· | ···················· | |
| L | 2957 | hCoV-19/USA/LA-EVTL251/2020\|EPI_ISL_574385\|2020-08 | ···················· | ···················· | ···················· | ···················· | ···················· | ···················· | |
| L | 2958 | hCoV-19/USA/LA-EVTL252/2020\|EPI_ISL_574386\|2020-08 | ···················· | ·A·················· | ···················· | ···················· | ···················· | ···················· | |
| L | 2959 | hCoV-19/USA/LA-EVTL253/2020\|EPI_ISL_574387\|2020-08 | ···················· | ···················· | ···················· | ···················· | ···················· | ···················· | |
| L | 2960 | hCoV-19/USA/LA-EVTL254/2020\|EPI_ISL_574388\|2020-08 | ···················· | ···················· | ···················· | ···················· | ···················· | ···················· | |
| L | 2961 | hCoV-19/USA/LA-EVTL258/2020\|EPI_ISL_574392\|2020-08 | ···················· | ···················· | ···················· | ···················· | ···················· | ···················· | |
| L | 2962 | hCoV-19/USA/LA-EVTL259/2020\|EPI_ISL_574393\|2020-08 | ···················· | ···················· | ···················· | ···················· | ···················· | ···················· | |
| L | 2963 | hCoV-19/USA/LA-EVTL260/2020\|EPI_ISL_574394\|2020-08 | ···················· | ···················· | ···················· | ···················· | ···················· | ···················· | |
| L | 2964 | hCoV-19/USA/LA-EVTL261/2020\|EPI_ISL_574395\|2020-08 | ···················· | ···················· | ···················· | ···················· | ···················· | ···················· | |
| L | 2965 | hCoV-19/England/ALDP-9CB572/2020\|EPI_ISL_575654\|20 | ···················· | ···················· | ···················· | ···················· | ···················· | ···················· | |
| L | 2966 | hCoV-19/England/ALDP-9CC0CB/2020\|EPI_ISL_575655\|20 | ···················· | ···················· | ···················· | ···················· | ···················· | ···················· | |
| L | 2967 | hCoV-19/England/ALDP-9CC034/2020\|EPI_ISL_575656\|20 | ···················· | ···················· | ···················· | ···················· | ···················· | ···················· | |
| L | 2968 | hCoV-19/England/ALDP-9CB396/2020\|EPI_ISL_575657\|20 | ···················· | ···················· | ···················· | ···················· | ···················· | ···················· | |
| L | 2969 | hCoV-19/England/ALDP-9CAD8A/2020\|EPI_ISL_575658\|20 | ···················· | ···················· | ···················· | ···················· | ···················· | ···················· | |
| L | 2970 | hCoV-19/England/ALDP-9CBF29/2020\|EPI_ISL_575660\|20 | ···················· | ···················· | ···················· | ···················· | ···················· | ···················· | |
| L | 2971 | hCoV-19/England/ALDP-9CB879/2020\|EPI_ISL_575661\|20 | ···················· | ···················· | ···················· | ···················· | ···················· | ···················· | |
| L | 2972 | hCoV-19/England/ALDP-9CBFFC/2020\|EPI_ISL_575662\|20 | ···················· | ···················· | ···················· | ···················· | ···················· | ···················· | |
| L | 2973 | hCoV-19/Wales/ALDP-9CBF83/2020\|EPI_ISL_575663\|2020 | ···················· | ···················· | ···················· | ···················· | ···················· | ···················· | |
| L | 2974 | hCoV-19/England/ALDP-9CBE59/2020\|EPI_ISL_575664\|20 | ···················· | ···················· | ···················· | ···················· | ···················· | ···················· | |
| L | 2975 | hCoV-19/England/ALDP-9CBD98/2020\|EPI_ISL_575665\|20 | ···················· | ···················· | ···················· | ···················· | ···················· | ···················· | |
| L | 2976 | hCoV-19/England/ALDP-9CAFFD/2020\|EPI_ISL_575666\|20 | ···················· | ···················· | ···················· | ···················· | ···················· | ···················· | |
| L | 2977 | hCoV-19/England/ALDP-9CBB61/2020\|EPI_ISL_575667\|20 | ···················· | ···················· | ···················· | ···················· | ···················· | ···················· | |
| L | 2978 | hCoV-19/England/ALDP-9CACD8/2020\|EPI_ISL_575668\|20 | ···················· | ···················· | ···················· | ···················· | ···················· | ···················· | |
| L | 2979 | hCoV-19/England/ALDP-9CC08F/2020\|EPI_ISL_575671\|20 | ···················· | ···················· | ···················· | ···················· | ···················· | ···················· | |
| L | 2980 | hCoV-19/England/ALDP-9CBC7D/2020\|EPI_ISL_575672\|20 | ···················· | ·A·······A·········· | ···················· | ···················· | ···················· | ···················· | |
| L | 2981 | hCoV-19/England/ALDP-9CBFCF/2020\|EPI_ISL_575673\|20 | ···················· | ···················· | ···················· | ···················· | ···················· | ···················· | |
| L | 2982 | hCoV-19/England/ALDP-9CAF39/2020\|EPI_ISL_575674\|20 | ···················· | ···················· | ···················· | ···················· | ···················· | ···················· | |
| L | 2983 | hCoV-19/England/ALDP-9CBF74/2020\|EPI_ISL_575675\|20 | ···················· | ···················· | ···················· | ···················· | ···················· | ···················· | |
| L | 2984 | hCoV-19/England/ALDP-9CB5DC/2020\|EPI_ISL_575676\|20 | ···················· | ···················· | ···················· | ···················· | ···················· | ···················· | |
| L | 2985 | hCoV-19/England/ALDP-9CBA73/2020\|EPI_ISL_575677\|20 | ··················T· | ···················· | ···················· | ···················· | ···················· | ···················· | 1 |
| L | 2986 | hCoV-19/England/ALDP-9CBB43/2020\|EPI_ISL_575678\|20 | ···················· | ···················· | ···················· | ···················· | ···················· | ···················· | |
| L | 2987 | hCoV-19/England/ALDP-9CAC32/2020\|EPI_ISL_575679\|20 | ···················· | ···················· | ···················· | ···················· | ···················· | ···················· | |
| L | 2988 | hCoV-19/England/ALDP-9CAE69/2020\|EPI_ISL_575680\|20 | ···················· | ···················· | ···················· | ···················· | ···················· | ···················· | |
| L | 2989 | hCoV-19/England/ALDP-9CBADD/2020\|EPI_ISL_575681\|20 | ···················· | ···················· | ···················· | ···················· | ···················· | ···················· | |
| L | 2990 | hCoV-19/England/ALDP-9CB26C/2020\|EPI_ISL_575682\|20 | ···················· | ···················· | ···················· | ···················· | ···················· | ···················· | |
| L | 2991 | hCoV-19/England/ALDP-9CAA0B/2020\|EPI_ISL_575684\|20 | ···················· | ·A·················· | ···················· | ···················· | ···················· | ···A················ | |

| | | | | | | | | |
|---|---|---|---|---|---|---|---|---|
| L | 2992 | hCoV-19/England/ALDP-9CB18D/2020\|EPI_ISL_575686\|20 | ···················· | ···················· | ····················· | ···················· | ···················· | ···················· |
| L | 2993 | hCoV-19/England/ALDP-9CBEB3/2020\|EPI_ISL_575687\|20 | ···················· | ···················· | ····················· | ···················· | ···················· | ···················· |
| L | 2994 | hCoV-19/England/ALDP-9CB75E/2020\|EPI_ISL_575688\|20 | ···················· | ···················· | ····················· | ···················· | ···················· | ···················· |
| L | 2995 | hCoV-19/England/ALDP-9CBE2C/2020\|EPI_ISL_575689\|20 | ···················· | ···················· | ····················· | ···················· | ···················· | ···················· |
| L | 2996 | hCoV-19/England/ALDP-9CB8C4/2020\|EPI_ISL_575691\|20 | ···················· | ·A·················· | ····················· | ···················· | ···················· | ···················· |
| L | 2997 | hCoV-19/England/ALDP-9CAA1A/2020\|EPI_ISL_575693\|20 | ···················· | ···················· | ····················· | ···················· | ···················· | ···················· |
| L | 2998 | hCoV-19/England/ALDP-9CB150/2020\|EPI_ISL_575694\|20 | ···················· | ···················· | ····················· | ···················· | ···················· | ···················· |
| L | 2999 | hCoV-19/England/ALDP-9CBF47/2020\|EPI_ISL_575695\|20 | ···················· | ···················· | ····················· | ···················· | ···················· | ···················· |
| L | 3000 | hCoV-19/England/ALDP-9CCCD6/2020\|EPI_ISL_575696\|20 | ···················· | ···················· | ····················· | ···················· | ···················· | ···················· |
| L | 3001 | hCoV-19/England/ALDP-9CB25D/2020\|EPI_ISL_575697\|20 | ···················· | ···················· | ····················· | ···················· | ···················· | ···················· |
| L | 3002 | hCoV-19/England/ALDP-9CAF75/2020\|EPI_ISL_575698\|20 | ···················· | ···················· | ····················· | ···················· | ···················· | ···················· |
| L | 3003 | hCoV-19/England/ALDP-9CB466/2020\|EPI_ISL_575699\|20 | ···················· | ···················· | ····················· | ···················· | ···················· | ···················· |
| L | 3004 | hCoV-19/England/LOND-D9923/2020\|EPI_ISL_576985\|202 | ···················· | ·············A······ | ····················· | ···················· | ···················· | ···················· |
| L | 3005 | hCoV-19/England/MILK-991715/2020\|EPI_ISL_576986\|20 | ···················· | ···················· | ····················· | ···················· | ···················· | ···················· |
| L | 3006 | hCoV-19/England/MILK-991733/2020\|EPI_ISL_576987\|20 | ···················· | ···················· | ····················· | ···················· | ···················· | ···················· |
| L | 3007 | hCoV-19/England/MILK-991742/2020\|EPI_ISL_576988\|20 | ···················· | ···················· | ····················· | ···················· | ···················· | ···················· |
| L | 3008 | hCoV-19/England/MILK-991751/2020\|EPI_ISL_576989\|20 | ···················· | ···················· | ····················· | ···················· | ···················· | ···················· |
| L | 3009 | hCoV-19/England/MILK-99177F/2020\|EPI_ISL_576991\|20 | ···················· | ···················· | ····················· | ···················· | ···················· | ···················· |
| L | 3010 | hCoV-19/England/MILK-9917AC/2020\|EPI_ISL_576992\|20 | ···················· | ···················· | ····················· | ···················· | ···················· | ···················· |
| L | 3012 | hCoV-19/England/MILK-9917D9/2020\|EPI_ISL_576994\|20 | ···················· | ···················· | ····················· | ···················· | ···················· | ···················· |
| L | 3013 | hCoV-19/England/MILK-991812/2020\|EPI_ISL_576995\|20 | ···················· | ···················· | ····················· | ···················· | ···················· | ···················· |
| L | 3014 | hCoV-19/England/MILK-99186D/2020\|EPI_ISL_576996\|20 | ···················· | ···················· | ····················· | ···················· | ···················· | ···················· |
| L | 3015 | hCoV-19/England/MILK-99187C/2020\|EPI_ISL_576997\|20 | ···················· | ···················· | ····················· | ···················· | ···················· | ···················· |
| L | 3016 | hCoV-19/England/MILK-99188B/2020\|EPI_ISL_576998\|20 | ···················· | ···················· | ····················· | ···················· | ···················· | ···················· |
| L | 3017 | hCoV-19/England/MILK-99189A/2020\|EPI_ISL_576999\|20 | ···················· | ···················· | ····················· | ···················· | ···················· | ···················· |
| L | 3018 | hCoV-19/New Zealand/20VR1904/2020\|EPI_ISL_579160\|2 | ···················· | ···················· | ····················· | ···················· | ···················· | ···················· |
| L | 3019 | hCoV-19/New Zealand/20VR1916/2020\|EPI_ISL_579163\|2 | ···················· | ···················· | ····················· | ···················· | ···················· | ···················· |
| L | 3020 | hCoV-19/New Zealand/20VR1923/2020\|EPI_ISL_579165\|2 | ···················· | ···················· | ····················· | ···················· | ···················· | ···················· |
| L | 3022 | hCoV-19/New Zealand/20VR1931/2020\|EPI_ISL_579167\|2 | ···················· | ···················· | ····················· | ···················· | ···················· | ···················· |
| L | 3024 | hCoV-19/New Zealand/20VR1940/2020\|EPI_ISL_579169\|2 | ···················· | ···················· | ····················· | ···················· | ···················· | ···················· |
| L | 3025 | hCoV-19/New Zealand/20VR1943/2020\|EPI_ISL_579170\|2 | ···················· | ···················· | ····················· | ···················· | ···················· | ···················· |
| L | 3026 | hCoV-19/New Zealand/20VR1944/2020\|EPI_ISL_579171\|2 | ···················· | ···················· | ····················· | ···················· | ···················· | ···················· |
| L | 3027 | hCoV-19/New Zealand/20VR1946/2020\|EPI_ISL_579173\|2 | ···················· | ···················· | ····················· | ···················· | ···················· | ···················· |
| L | 3028 | hCoV-19/New Zealand/20VR1948/2020\|EPI_ISL_579174\|2 | ···················· | ···················· | ····················· | ···················· | ···················· | ···················· |
| L | 3030 | hCoV-19/New Zealand/20VR1956/2020\|EPI_ISL_579176\|2 | ···················· | ···················· | ····················· | ···················· | ···················· | ···················· |
| L | 3031 | hCoV-19/New Zealand/20VR1957/2020\|EPI_ISL_579177\|2 | ···················· | ···················· | ····················· | ···················· | ···················· | ···················· |
| L | 3032 | hCoV-19/New Zealand/20VR1959/2020\|EPI_ISL_579178\|2 | ···················· | ···················· | ····················· | ···················· | ···················· | ···················· |
| L | 3033 | hCoV-19/New Zealand/20VR1960/2020\|EPI_ISL_579179\|2 | ···················· | ···················· | ····················· | ···················· | ···················· | ···················· |
| L | 3034 | hCoV-19/England/ALDP-9EA373/2020\|EPI_ISL_580120\|20 | ···················· | ···················· | ····················· | ···················· | ···················· | ···················· |
| L | 3035 | hCoV-19/England/ALDP-9E76E6/2020\|EPI_ISL_580121\|20 | ···················· | ·A·················· | ····················· | ···················· | ···················· | ···················· |
| L | 3036 | hCoV-19/England/ALDP-9EA7A4/2020\|EPI_ISL_580122\|20 | ···················· | ···················· | ····················· | ···················· | ···················· | ···················· |
| L | 3037 | hCoV-19/England/ALDP-9EA0E5/2020\|EPI_ISL_580123\|20 | ···················· | ···················· | ····················· | ···················· | ···················· | ···················· |
| L | 3038 | hCoV-19/England/ALDP-9EA980/2020\|EPI_ISL_580124\|20 | ···················· | ···················· | ····················· | ···················· | ···················· | ···················· |
| L | 3039 | hCoV-19/England/ALDP-9EB0B7/2020\|EPI_ISL_580125\|20 | ···················· | ···················· | ····················· | ···················· | ···················· | ···················· |
| L | 3040 | hCoV-19/England/ALDP-9EAF9D/2020\|EPI_ISL_580127\|20 | ···················· | ···················· | ····················· | ···················· | ···················· | ···················· |
| L | 3041 | hCoV-19/England/ALDP-9E9E55/2020\|EPI_ISL_580128\|20 | ···················· | ···················· | ····················· | ···················· | ···················· | ···················· |
| L | 3042 | hCoV-19/England/ALDP-9EA2A3/2020\|EPI_ISL_580129\|20 | ···················· | ···················· | ····················· | ···················· | ···················· | ···················· |
| L | 3043 | hCoV-19/England/ALDP-9EA0C7/2020\|EPI_ISL_580130\|20 | ···················· | ···················· | ····················· | ···················· | ···················· | ···················· |
| L | 3044 | hCoV-19/England/ALDP-9EA908/2020\|EPI_ISL_580131\|20 | ···················· | ···················· | ····················· | ···················· | ···················· | ···················· |
| L | 3045 | hCoV-19/England/ALDP-9E9CC4/2020\|EPI_ISL_580133\|20 | ···················· | ···················· | ····················· | ···················· | ···················· | ···················· |
| L | 3046 | hCoV-19/England/ALDP-9EA8ED/2020\|EPI_ISL_580135\|20 | ···················· | ···················· | ····················· | ···················· | ···················· | ···················· |
| L | 3047 | hCoV-19/England/ALDP-9EAD75/2020\|EPI_ISL_580136\|20 | ···················· | ···················· | ····················· | ···················· | ···················· | ···················· |
| L | 3048 | hCoV-19/England/ALDP-9E9E28/2020\|EPI_ISL_580139\|20 | ···················· | ···················· | ····················· | ···················· | ···················· | ···················· |
| L | 3049 | hCoV-19/England/ALDP-9E9C79/2020\|EPI_ISL_580141\|20 | ···················· | ···················· | ····················· | ···················· | ···················· | ···················· |
| L | 3050 | hCoV-19/England/MILK-970460/2020\|EPI_ISL_581401\|20 | ···················· | ·A·················· | ····················· | ···················· | ···················· | ···A················ |
| L | 3051 | hCoV-19/England/QEUH-96E81E/2020\|EPI_ISL_581402\|20 | ···················· | ···················· | ····················· | ···················· | ···················· | ···················· |
| L | 3052 | hCoV-19/England/QEUH-96A029/2020\|EPI_ISL_581403\|20 | ···················· | ···················· | ····················· | ···················· | ···················· | ···················· |
| L | 3053 | hCoV-19/England/QEUH-964B6A/2020\|EPI_ISL_581405\|20 | ···················· | ·A·················· | ····················· | ···················· | ···················· | ···················· |
| L | 3054 | hCoV-19/England/QEUH-96BC33/2020\|EPI_ISL_581407\|20 | ···················· | ···················· | ····················· | ···················· | ···················· | ···················· |
| L | 3055 | hCoV-19/England/QEUH-96B091/2020\|EPI_ISL_581408\|20 | ···················· | ···················· | ····················· | ···················· | ···················· | ···················· |
| L | 3056 | hCoV-19/England/ALDP-9545AB/2020\|EPI_ISL_581409\|20 | ···················· | ···················· | ····················· | ···················· | ···················· | ···················· |
| L | 3057 | hCoV-19/India/WB-IICB-019/2020\|EPI_ISL_581451\|2020 | ···················· | ···················· | ····················· | ···················· | ···················· | ···················· |
| L | 3058 | hCoV-19/Sweden/20-21509/2020\|EPI_ISL_582782\|2020-0 | ···················· | ···················· | ····················· | ···················· | ···················· | ···················· |
| L | 3059 | hCoV-19/Sweden/20-21512/2020\|EPI_ISL_582783\|2020-0 | ···················· | ···················· | ····················· | ···················· | ···················· | ···················· |
| L | 3060 | hCoV-19/Sweden/20-21542/2020\|EPI_ISL_582784\|2020-0 | ···················· | ···················· | ····················· | ···················· | ···················· | ···················· |
| L | 3061 | hCoV-19/Sweden/20-22107/2020\|EPI_ISL_582785\|2020-0 | ···················· | ···················· | ····················· | ···················· | ···················· | ···················· |
| L | 3062 | hCoV-19/Sweden/20-22111/2020\|EPI_ISL_582786\|2020-0 | ···················· | ···················· | ····················· | ···················· | ···················· | ···················· |
| L | 3063 | hCoV-19/Sweden/20-22112/2020\|EPI_ISL_582787\|2020-0 | ···················· | ···················· | ····················· | ···················· | ···················· | ···················· |
| L | 3064 | hCoV-19/Sweden/20-22122/2020\|EPI_ISL_582791\|2020-0 | ···················· | ····G··············· | ····················· | ···················· | ···················· | ···················· |
| L | 3065 | hCoV-19/Sweden/20-22125/2020\|EPI_ISL_582792\|2020-0 | ···················· | ···················· | ····················· | ···················· | ···················· | ···················· |
| L | 3066 | hCoV-19/Sweden/20-22126/2020\|EPI_ISL_582793\|2020-0 | ···················· | ···················· | ····················· | ···················· | ···················· | ···················· |
| L | 3067 | hCoV-19/Sweden/20-51972/2020\|EPI_ISL_582794\|2020-0 | ···················· | ···················· | ····················· | ···················· | ···················· | ···················· |
| L | 3068 | hCoV-19/Sweden/20-51974/2020\|EPI_ISL_582795\|2020-0 | ···················· | ···················· | ····················· | ···················· | ···················· | ···················· |
| L | 3069 | hCoV-19/Sweden/20-51975/2020\|EPI_ISL_582796\|2020-0 | ···················· | ···················· | ····················· | ···················· | ···················· | ···················· |
| L | 3070 | hCoV-19/Sweden/20-52269/2020\|EPI_ISL_582797\|2020-0 | ···················· | ···················· | ····················· | ···················· | ···················· | ···················· |
| L | 3071 | hCoV-19/Sweden/20-52276/2020\|EPI_ISL_582798\|2020-0 | ···················· | ···················· | ····················· | ···················· | ···················· | ···················· |
| L | 3072 | hCoV-19/Sweden/20-52282/2020\|EPI_ISL_582799\|2020-0 | ···················· | ···················· | ····················· | ···················· | ···················· | ···················· |
| L | 3073 | hCoV-19/USA/NJ-QDX-1209/2020\|EPI_ISL_571970\|2020-0 | ···················· | ···················· | ····················· | ···················· | ···················· | ···················· |
| L | 3074 | hCoV-19/USA/NJ-QDX-1210/2020\|EPI_ISL_571971\|2020-0 | ···················· | ···················· | ····················· | ···················· | ···················· | ···················· |
| L | 3075 | hCoV-19/USA/NJ-QDX-1211/2020\|EPI_ISL_571972\|2020-0 | ···················· | ···················· | ····················· | ···················· | ···················· | ···················· |

| | | | | | | | | |
|---|---|---|---|---|---|---|---|---|
| L | 3076 | hCoV-19/USA/NJ-QDX-1212/2020\|EPI_ISL_571973\|2020-0 | ···················· | ···················· | ···················· | ···················· | ···················· | ···················· |
| L | 3077 | hCoV-19/USA/NJ-QDX-1213/2020\|EPI_ISL_571974\|2020-0 | ···················· | ···················· | ···················· | ···················· | ···················· | ···················· |
| L | 3078 | hCoV-19/USA/NJ-QDX-1216/2020\|EPI_ISL_571975\|2020-0 | ···················· | ···················· | ···················· | ···················· | ···················· | ···················· |
| L | 3079 | hCoV-19/USA/NJ-QDX-1217/2020\|EPI_ISL_571976\|2020-0 | ···················· | ···················· | ···················· | ···················· | ···················· | ···················· |
| L | 3080 | hCoV-19/USA/NJ-QDX-1221/2020\|EPI_ISL_571977\|2020-0 | ···················· | ···················· | ···················· | ···················· | ···················· | ···················· |
| L | 3081 | hCoV-19/USA/NJ-QDX-1223/2020\|EPI_ISL_571978\|2020-0 | ···················· | ···················· | ···················· | ···················· | ···················· | ···················· |
| L | 3082 | hCoV-19/USA/NJ-QDX-1252/2020\|EPI_ISL_571979\|2020-0 | ···················· | ···················· | ···················· | ···················· | ···················· | ···················· |
| L | 3083 | hCoV-19/USA/NJ-QDX-1253/2020\|EPI_ISL_571980\|2020-0 | ···················· | ···················· | ···················· | ···················· | ···················· | ···················· |
| L | 3084 | hCoV-19/USA/NJ-QDX-1254/2020\|EPI_ISL_571981\|2020-0 | ···················· | ···················· | ···················· | ···················· | ···················· | ···················· |
| L | 3085 | hCoV-19/USA/NJ-QDX-1267/2020\|EPI_ISL_571982\|2020-0 | ···················· | ···················· | ···················· | ···················· | ···················· | ···················· |
| L | 3086 | hCoV-19/USA/NJ-QDX-1269/2020\|EPI_ISL_571983\|2020-0 | ···················· | ···················· | ···················· | ···················· | ···················· | ···················· |
| L | 3087 | hCoV-19/USA/NY-QDX-1174/2020\|EPI_ISL_571984\|2020-0 | ···················· | ···················· | ···················· | ···················· | ···················· | ···················· |
| L | 3088 | hCoV-19/USA/NY-QDX-1175/2020\|EPI_ISL_571985\|2020-0 | ···················· | ···················· | ···················· | ···················· | ···················· | ···················· |
| L | 3089 | hCoV-19/USA/NY-QDX-1176/2020\|EPI_ISL_571986\|2020-0 | ···················· | ···················· | ···················· | ···················· | ···················· | ···················· |
| L | 3090 | hCoV-19/USA/NY-QDX-1177/2020\|EPI_ISL_571987\|2020-0 | ···················· | ···················· | ···················· | ···················· | ···················· | ···················· |
| L | 3091 | hCoV-19/USA/NY-QDX-1178/2020\|EPI_ISL_571988\|2020-0 | ···················· | ···················· | ···················· | ···················· | ···················· | ···················· |
| L | 3092 | hCoV-19/USA/NY-QDX-1179/2020\|EPI_ISL_571989\|2020-0 | ···················· | ···················· | ···················· | ···················· | ···················· | ···················· |
| L | 3093 | hCoV-19/USA/NY-QDX-1180/2020\|EPI_ISL_571990\|2020-0 | ···················· | ···················· | ···················· | ···················· | ···················· | ···················· |
| L | 3094 | hCoV-19/England/ALDP-9BCBAF/2020\|EPI_ISL_573058\|20 | ···················· | ·A·················· | ···················· | ···················· | ···················· | ··A················· |
| L | 3095 | hCoV-19/England/ALDP-9BCBBE/2020\|EPI_ISL_573059\|20 | ···················· | ···················· | ···················· | ···················· | ···················· | ···················· |
| L | 3096 | hCoV-19/England/ALDP-9BCBCD/2020\|EPI_ISL_573060\|20 | ···················· | ···················· | ···················· | ···················· | ···················· | ···················· |
| L | 3097 | hCoV-19/England/ALDP-9BCBEB/2020\|EPI_ISL_573062\|20 | ···················· | ···················· | ···················· | ···················· | ···················· | ···················· |
| L | 3098 | hCoV-19/England/ALDP-9BCC15/2020\|EPI_ISL_573065\|20 | ···················· | ·A·················· | ···················· | ···················· | ···················· | ··A················· |
| L | 3099 | hCoV-19/England/ALDP-9BCC24/2020\|EPI_ISL_573066\|20 | ···················· | ···················· | ···················· | ···················· | ···················· | ···················· |
| L | 3100 | hCoV-19/England/ALDP-9BCC8E/2020\|EPI_ISL_573067\|20 | ···················· | ···················· | ···················· | ···················· | ···················· | ···················· |
| L | 3101 | hCoV-19/England/ALDP-9BCC9D/2020\|EPI_ISL_573068\|20 | ···················· | ···················· | ···················· | ···················· | ···················· | ···················· |
| L | 3102 | hCoV-19/USA/LA-EVTL165/2020\|EPI_ISL_574300\|2020-05 | ···················· | ···················· | ···················· | ···················· | ···················· | ···················· |
| L | 3103 | hCoV-19/USA/LA-EVTL166/2020\|EPI_ISL_574301\|2020-05 | ···················· | ···················· | ···················· | ···················· | ···················· | ···················· |
| L | 3104 | hCoV-19/USA/LA-EVTL167/2020\|EPI_ISL_574302\|2020-05 | ···················· | ···················· | ···················· | ···················· | ···················· | ···················· |
| L | 3105 | hCoV-19/USA/LA-EVTL168/2020\|EPI_ISL_574303\|2020-05 | ···················· | ···················· | ···················· | ···················· | ···················· | ···················· |
| L | 3106 | hCoV-19/USA/LA-EVTL169/2020\|EPI_ISL_574304\|2020-05 | ···················· | ···················· | ···················· | ···················· | ···················· | ···················· |
| L | 3107 | hCoV-19/USA/LA-EVTL170/2020\|EPI_ISL_574305\|2020-05 | ···················· | ···················· | ···················· | ···················· | ···················· | ···················· |
| L | 3108 | hCoV-19/USA/LA-EVTL171/2020\|EPI_ISL_574306\|2020-05 | ···················· | ···················· | ···················· | ···················· | ···················· | ···················· |
| L | 3109 | hCoV-19/USA/LA-EVTL172/2020\|EPI_ISL_574307\|2020-05 | ···················· | ···················· | ···················· | ···················· | ···················· | ···················· |
| L | 3110 | hCoV-19/USA/LA-EVTL173/2020\|EPI_ISL_574308\|2020-05 | ···················· | ···················· | ···················· | ···················· | ···················· | ···················· |
| L | 3111 | hCoV-19/USA/LA-EVTL174/2020\|EPI_ISL_574309\|2020-05 | ···················· | ···················· | ···················· | ···················· | ···················· | ···················· |
| L | 3112 | hCoV-19/USA/LA-EVTL175/2020\|EPI_ISL_574310\|2020-05 | ···················· | ···················· | ···················· | ···················· | ···················· | ···················· |
| L | 3113 | hCoV-19/USA/LA-EVTL176/2020\|EPI_ISL_574311\|2020-05 | ···················· | ···················· | ···················· | ···················· | ···················· | ···················· |
| L | 3114 | hCoV-19/USA/LA-EVTL177/2020\|EPI_ISL_574312\|2020-05 | ···················· | ···················· | ···················· | ···················· | ···················· | ···················· |
| L | 3115 | hCoV-19/USA/LA-EVTL178/2020\|EPI_ISL_574313\|2020-05 | ···················· | ···················· | ···················· | ···················· | ···················· | ···················· |
| L | 3116 | hCoV-19/USA/LA-EVTL179/2020\|EPI_ISL_574314\|2020-05 | ···················· | ···················· | ···················· | ···················· | ···················· | ···················· |
| L | 3117 | hCoV-19/USA/LA-EVTL180/2020\|EPI_ISL_574315\|2020-05 | ···················· | ···················· | ···················· | ···················· | ···················· | ···················· |
| L | 3118 | hCoV-19/USA/LA-EVTL181/2020\|EPI_ISL_574316\|2020-05 | ···················· | ···················· | ···················· | ···················· | ···················· | ···················· |
| L | 3119 | hCoV-19/USA/LA-EVTL182/2020\|EPI_ISL_574317\|2020-07 | ···················· | ···················· | ···················· | ···················· | ···················· | ···················· |
| L | 3120 | hCoV-19/USA/LA-EVTL183/2020\|EPI_ISL_574318\|2020-07 | ···················· | ···················· | ···················· | ···················· | ···················· | ···················· |
| L | 3121 | hCoV-19/USA/LA-EVTL184/2020\|EPI_ISL_574319\|2020-07 | ···················· | ···················· | ···················· | ···················· | ···················· | ···················· |
| L | 3122 | hCoV-19/USA/LA-EVTL186/2020\|EPI_ISL_574320\|2020-05 | ···················· | ···················· | ···················· | ···················· | ···················· | ···················· |
| L | 3123 | hCoV-19/USA/LA-EVTL187/2020\|EPI_ISL_574321\|2020-05 | ···················· | ···················· | ···················· | ···················· | ···················· | ···················· |
| L | 3124 | hCoV-19/USA/LA-EVTL188/2020\|EPI_ISL_574322\|2020-05 | ···················· | ···················· | ···················· | ···················· | ···················· | ···················· |
| L | 3125 | hCoV-19/USA/LA-EVTL255/2020\|EPI_ISL_574389\|2020-08 | ···················· | ···················· | ···················· | ···················· | ···················· | ···················· |
| L | 3126 | hCoV-19/USA/LA-EVTL256/2020\|EPI_ISL_574390\|2020-08 | ···················· | ···················· | ···················· | ···················· | ···················· | ···················· |
| L | 3127 | hCoV-19/USA/LA-EVTL257/2020\|EPI_ISL_574391\|2020-08 | ···················· | ···················· | ···················· | ···················· | ···················· | ···················· |
| L | 3128 | hCoV-19/USA/LA-EVTL262/2020\|EPI_ISL_574396\|2020-08 | ···················· | ···················· | ···················· | ···················· | ···················· | ···················· |
| L | 3129 | hCoV-19/USA/LA-EVTL263/2020\|EPI_ISL_574397\|2020-08 | ···················· | ···················· | ···················· | ···················· | ···················· | ···················· |
| L | 3130 | hCoV-19/USA/LA-EVTL264/2020\|EPI_ISL_574398\|2020-08 | ···················· | ·A·················· | ···················· | ···················· | ···················· | ···················· |
| L | 3131 | hCoV-19/USA/LA-EVTL265/2020\|EPI_ISL_574399\|2020-08 | ···················· | ···················· | ···················· | ···················· | ···················· | ···················· |
| L | 3132 | hCoV-19/England/ALDP-9CB518/2020\|EPI_ISL_575600\|20 | ···················· | ···················· | ···················· | ···················· | ···················· | ···················· |
| L | 3133 | hCoV-19/England/ALDP-9CBAFB/2020\|EPI_ISL_575601\|20 | ···················· | ···················· | ···················· | ···················· | ···················· | ···················· |
| L | 3134 | hCoV-19/England/ALDP-9CB105/2020\|EPI_ISL_575602\|20 | ···················· | ···················· | ···················· | ···················· | ···················· | ···················· |
| L | 3135 | hCoV-19/England/ALDP-9CBCC8/2020\|EPI_ISL_575603\|20 | ···················· | ···················· | ···················· | ···················· | ···················· | ···················· |
| L | 3136 | hCoV-19/England/ALDP-9CB33C/2020\|EPI_ISL_575604\|20 | ···················· | ···················· | ···················· | ···················· | ···················· | ···················· |
| L | 3137 | hCoV-19/England/ALDP-9CB6E8/2020\|EPI_ISL_575605\|20 | ···················· | ···················· | ···················· | ···················· | ···················· | ···················· |
| L | 3138 | hCoV-19/England/ALDP-9CAD99/2020\|EPI_ISL_575606\|20 | ···················· | ···················· | ···················· | ···················· | ···················· | ···················· |
| L | 3139 | hCoV-19/England/ALDP-9CB9D0/2020\|EPI_ISL_575607\|20 | ···················· | ···················· | ···················· | ···················· | ···················· | ···················· |
| L | 3140 | hCoV-19/England/ALDP-9CB1E7/2020\|EPI_ISL_575609\|20 | ···················· | ·A·················· | ···················· | ···················· | ···················· | ··A················· |
| L | 3141 | hCoV-19/England/ALDP-9CB2E4/2020\|EPI_ISL_575610\|20 | ···················· | ···················· | ···················· | ···················· | ···················· | ···················· |
| L | 3142 | hCoV-19/England/ALDP-9CB9A3/2020\|EPI_ISL_575611\|20 | ···················· | ···················· | ···················· | ···················· | ···················· | ···················· |
| L | 3143 | hCoV-19/England/ALDP-9CB74F/2020\|EPI_ISL_575612\|20 | ···················· | ···················· | ···················· | ···················· | ···················· | ···················· |
| L | 3144 | hCoV-19/England/ALDP-9CBDB6/2020\|EPI_ISL_575614\|20 | ···················· | ···················· | ···················· | ···················· | ···················· | ···················· |
| L | 3145 | hCoV-19/England/ALDP-9CBE95/2020\|EPI_ISL_575615\|20 | ···················· | ···················· | ···················· | ···················· | ···················· | ···················· |
| L | 3146 | hCoV-19/England/ALDP-9CAED2/2020\|EPI_ISL_575616\|20 | ···················· | ···················· | ···················· | ···················· | ···················· | ···················· |
| L | 3147 | hCoV-19/England/ALDP-9CB31E/2020\|EPI_ISL_575617\|20 | ···················· | ···················· | ···················· | ···················· | ···················· | ···················· |
| L | 3148 | hCoV-19/England/ALDP-9CAD6C/2020\|EPI_ISL_575618\|20 | ···················· | ···················· | ···················· | ···················· | ···················· | ···················· |
| L | 3149 | hCoV-19/England/ALDP-9CB509/2020\|EPI_ISL_575619\|20 | ···················· | ···················· | ···················· | ···················· | ···················· | ···················· |
| L | 3150 | hCoV-19/England/ALDP-9CB78B/2020\|EPI_ISL_575620\|20 | ···················· | ···················· | ···················· | ···················· | ···················· | ···················· |
| L | 3151 | hCoV-19/England/ALDP-9CB85B/2020\|EPI_ISL_575621\|20 | ···················· | ···················· | ···················· | ···················· | ···················· | ···················· |
| L | 3152 | hCoV-19/England/ALDP-9CB1AB/2020\|EPI_ISL_575622\|20 | ···················· | ···················· | ···················· | ···················· | ···················· | ···················· |
| L | 3153 | hCoV-19/England/ALDP-9CB0F9/2020\|EPI_ISL_575623\|20 | ···················· | ···················· | ···················· | ···················· | ···················· | ···················· |
| L | 3154 | hCoV-19/England/ALDP-9CB5BE/2020\|EPI_ISL_575624\|20 | ···················· | ···················· | ···················· | ···················· | ···················· | ···················· |
| L | 3155 | hCoV-19/England/ALDP-9CAB53/2020\|EPI_ISL_575625\|20 | ···················· | ···················· | ···················· | ···················· | ···················· | ···················· |

| | | | | | | | | |
|---|---|---|---|---|---|---|---|---|
| L | 3156 | hCoV-19/England/ALDP-9CB4A2/2020\|EPI_ISL_575626\|20 | ···················· | ···················· | ···················· | ···················· | ···················· | ···················· |
| L | 3157 | hCoV-19/England/ALDP-9CB730/2020\|EPI_ISL_575627\|20 | ···················· | ···················· | ···················· | ···················· | ···················· | ···················· |
| L | 3158 | hCoV-19/England/ALDP-9CBAEC/2020\|EPI_ISL_575628\|20 | ···················· | ···················· | ···················· | ···················· | ···················· | ···················· |
| L | 3159 | hCoV-19/England/ALDP-9CB5CD/2020\|EPI_ISL_575629\|20 | ···················· | ···················· | ···················· | ···················· | ···················· | ···················· |
| L | 3160 | hCoV-19/England/ALDP-9CACC9/2020\|EPI_ISL_575630\|20 | ···················· | ···················· | ···················· | ···················· | ···················· | ···················· |
| L | 3161 | hCoV-19/England/ALDP-9CC061/2020\|EPI_ISL_575631\|20 | ···················· | ···················· | ···················· | ···················· | ···················· | ···················· |
| L | 3162 | hCoV-19/England/ALDP-9CB0EA/2020\|EPI_ISL_575632\|20 | ···················· | ···················· | ···················· | ···················· | ···················· | ···················· |
| L | 3163 | hCoV-19/England/ALDP-9CAB44/2020\|EPI_ISL_575633\|20 | ···················· | ···················· | ···················· | ···················· | ···················· | ···················· |
| L | 3164 | hCoV-19/England/ALDP-9CB77C/2020\|EPI_ISL_575634\|20 | ···················· | ···················· | ···················· | ···················· | ···················· | ···················· |
| L | 3165 | hCoV-19/England/ALDP-9CB642/2020\|EPI_ISL_575635\|20 | ···················· | ···················· | ···················· | ···················· | ···················· | ···················· |
| L | 3166 | hCoV-19/England/ALDP-9CB16F/2020\|EPI_ISL_575636\|20 | ···················· | ·A·················· | ···················· | ···················· | ···················· | ··A················· |
| L | 3167 | hCoV-19/England/ALDP-9CBC04/2020\|EPI_ISL_575637\|20 | ···················· | ···················· | ···················· | ···················· | ···················· | ···················· |
| L | 3168 | hCoV-19/England/ALDP-9CB17E/2020\|EPI_ISL_575638\|20 | ···················· | ···················· | ···················· | ···················· | ···················· | ···················· |
| L | 3169 | hCoV-19/England/ALDP-9CAFC0/2020\|EPI_ISL_575639\|20 | ···················· | ···················· | ···················· | ···················· | ···················· | ···················· |
| L | 3170 | hCoV-19/England/ALDP-9CBC5F/2020\|EPI_ISL_575640\|20 | ···················· | ···················· | ···················· | ···················· | ···················· | ···················· |
| L | 3171 | hCoV-19/England/ALDP-9CBACE/2020\|EPI_ISL_575641\|20 | ···················· | ·A·················· | ···················· | ···················· | ···················· | ··A················· |
| L | 3172 | hCoV-19/England/ALDP-9CB114/2020\|EPI_ISL_575642\|20 | ···················· | ···················· | ···················· | ···················· | ···················· | ···················· |
| L | 3173 | hCoV-19/England/ALDP-9CBBCB/2020\|EPI_ISL_575643\|20 | ···················· | ···················· | ···················· | ···················· | ···················· | ···················· |
| L | 3174 | hCoV-19/England/ALDP-9CA9D1/2020\|EPI_ISL_575644\|20 | ···················· | ···················· | ···················· | ···················· | ···················· | ···················· |
| L | 3175 | hCoV-19/England/ALDP-9CBD01/2020\|EPI_ISL_575645\|20 | ···················· | ···················· | ···················· | ···················· | ···················· | ···················· |
| L | 3176 | hCoV-19/England/ALDP-9CBBBC/2020\|EPI_ISL_575646\|20 | ···················· | ···················· | ···················· | ···················· | ···················· | ···················· |
| L | 3177 | hCoV-19/England/ALDP-9CBEC2/2020\|EPI_ISL_575647\|20 | ···················· | ···················· | ···················· | ···················· | ···················· | ···················· |
| L | 3178 | hCoV-19/England/ALDP-9CB68E/2020\|EPI_ISL_575648\|20 | ···················· | ···················· | ···················· | ···················· | ···················· | ···················· |
| L | 3179 | hCoV-19/England/ALDP-9CBA46/2020\|EPI_ISL_575649\|20 | ···················· | ···················· | ···················· | ···················· | ···T················ | ···················· |
| L | 3180 | hCoV-19/England/ALDP-9CB606/2020\|EPI_ISL_575650\|20 | ···················· | ···················· | ···················· | ···················· | ···················· | ···················· |
| L | 3181 | hCoV-19/England/ALDP-9CB369/2020\|EPI_ISL_575651\|20 | ···················· | ···················· | ···················· | ···················· | ···················· | ···················· |
| L | 3182 | hCoV-19/England/ALDP-9CBB25/2020\|EPI_ISL_575652\|20 | ···················· | ···················· | ···················· | ···················· | ···················· | ···················· |
| L | 3183 | hCoV-19/England/ALDP-9CC0AD/2020\|EPI_ISL_575653\|20 | ···················· | ···················· | ···················· | ···················· | ···················· | ···················· |
| L | 3184 | hCoV-19/England/CAMB-1B634B/2020\|EPI_ISL_576900\|20 | ···················· | ···················· | ···················· | ···················· | ···················· | ···················· |
| L | 3185 | hCoV-19/England/CAMB-1B6369/2020\|EPI_ISL_576901\|20 | ···················· | ···················· | ···················· | ···················· | ···················· | ···················· |
| L | 3186 | hCoV-19/England/CAMB-1B63D2/2020\|EPI_ISL_576903\|20 | ···················· | ·A·················· | ···················· | ···················· | ···T················ | ··A················· |
| L | 3187 | hCoV-19/England/CAMB-1B641B/2020\|EPI_ISL_576906\|20 | ···················· | ···················· | ···················· | ···················· | ···················· | ···················· |
| L | 3188 | hCoV-19/England/CAMB-1B6439/2020\|EPI_ISL_576908\|20 | ···················· | ···················· | ···················· | ···················· | ···················· | ···················· |
| L | 3189 | hCoV-19/England/CAMB-1B6457/2020\|EPI_ISL_576910\|20 | ···················· | ···················· | ···················· | ···················· | ···················· | ···················· |
| L | 3191 | hCoV-19/England/CAMB-1B64B1/2020\|EPI_ISL_576914\|20 | ···················· | ···················· | ···················· | ···················· | ···················· | ···················· |
| L | 3192 | hCoV-19/England/CAMB-1B6509/2020\|EPI_ISL_576918\|20 | ···················· | ···················· | ···················· | ···················· | ···················· | ···················· |
| L | 3193 | hCoV-19/England/CAMC-9DF97F/2020\|EPI_ISL_576920\|20 | ···················· | ···················· | ···················· | ···················· | ···················· | ···················· |
| L | 3194 | hCoV-19/England/CAMC-9DF9D9/2020\|EPI_ISL_576921\|20 | ···················· | ·A·················· | ···················· | ···················· | ···················· | ··A················· |
| L | 3195 | hCoV-19/England/CAMC-9DFA03/2020\|EPI_ISL_576922\|20 | ···················· | ···················· | ···················· | ···················· | ···················· | ···················· |
| L | 3196 | hCoV-19/England/CAMC-9DFB1F/2020\|EPI_ISL_576923\|20 | ···················· | ···················· | ···················· | ···················· | ···················· | ···················· |
| L | 3197 | hCoV-19/England/CAMC-9DFBB5/2020\|EPI_ISL_576924\|20 | ···················· | ···················· | ···················· | ···················· | ···················· | ···················· |
| L | 3198 | hCoV-19/England/CAMC-9DFBC4/2020\|EPI_ISL_576925\|20 | ···················· | ···················· | ···················· | ···················· | ···················· | ···················· |
| L | 3199 | hCoV-19/England/CAMC-9DFBD3/2020\|EPI_ISL_576926\|20 | ···················· | ···················· | ···················· | ···················· | ···················· | ···················· |
| L | 3200 | hCoV-19/England/CAMC-9DFC1C/2020\|EPI_ISL_576927\|20 | ···················· | ···················· | ···················· | ···················· | ···················· | ···················· |
| L | 3201 | hCoV-19/England/CAMC-9DFC85/2020\|EPI_ISL_576928\|20 | ···················· | ···················· | ···················· | ···················· | ···················· | ···················· |
| L | 3202 | hCoV-19/England/CAMC-9DFCA3/2020\|EPI_ISL_576930\|20 | ···················· | ···················· | ···················· | ···················· | ···················· | ···················· |
| L | 3203 | hCoV-19/England/CAMC-9DFCB2/2020\|EPI_ISL_576931\|20 | ···················· | ···················· | ···················· | ···················· | ···················· | ···················· |
| L | 3204 | hCoV-19/England/CAMC-9DFCD0/2020\|EPI_ISL_576932\|20 | ···················· | ···················· | ···················· | ···················· | ···················· | ···················· |
| L | 3205 | hCoV-19/England/CAMC-9DFCEF/2020\|EPI_ISL_576933\|20 | ···················· | ·····A·············· | ···················· | ···················· | ···················· | ···················· |
| L | 3206 | hCoV-19/England/CAMC-9DFCFE/2020\|EPI_ISL_576934\|20 | ···················· | ···················· | ···················· | ···················· | ···················· | ···················· |
| L | 3207 | hCoV-19/England/CAMC-9DFD19/2020\|EPI_ISL_576935\|20 | ···················· | ···················· | ···················· | ···················· | ···················· | ···················· |
| L | 3208 | hCoV-19/England/CAMC-9DFD64/2020\|EPI_ISL_576936\|20 | ···················· | ···················· | ···················· | ···················· | ···················· | ···················· |
| L | 3209 | hCoV-19/England/CAMC-9DFD73/2020\|EPI_ISL_576937\|20 | ···················· | ···················· | ···················· | ···················· | ···················· | ···················· |
| L | 3210 | hCoV-19/England/CAMC-9DFDCE/2020\|EPI_ISL_576938\|20 | ···················· | ···················· | ···················· | ···················· | ···················· | ···················· |
| L | 3211 | hCoV-19/England/CAMC-9DFE34/2020\|EPI_ISL_576939\|20 | ···················· | ···················· | ···················· | ···················· | ···················· | ···················· |
| L | 3212 | hCoV-19/England/CAMC-9DFE61/2020\|EPI_ISL_576940\|20 | ···················· | ···················· | ···················· | ···················· | ···················· | ···················· |
| L | 3213 | hCoV-19/England/CAMC-9DFE9E/2020\|EPI_ISL_576941\|20 | ···················· | ···················· | ···················· | ···················· | ···················· | ···················· |
| L | 3214 | hCoV-19/England/CAMC-9DFECB/2020\|EPI_ISL_576942\|20 | ···················· | ···················· | ···················· | ···················· | ···················· | ···················· |
| L | 3215 | hCoV-19/England/CAMC-9DFEE9/2020\|EPI_ISL_576943\|20 | ···················· | ···················· | ···················· | ···················· | ···················· | ···················· |
| L | 3216 | hCoV-19/England/CAMC-9DFEF8/2020\|EPI_ISL_576944\|20 | ···················· | ···················· | ···················· | ···················· | ···················· | ···················· |
| L | 3217 | hCoV-19/England/CAMC-9DFF6E/2020\|EPI_ISL_576946\|20 | ···················· | ···················· | ···················· | ···················· | ···················· | ···················· |
| L | 3218 | hCoV-19/England/CAMC-9DFF9B/2020\|EPI_ISL_576947\|20 | ···················· | ···················· | ···················· | ···················· | ···················· | ···················· |
| L | 3219 | hCoV-19/England/CAMC-9DFFAA/2020\|EPI_ISL_576948\|20 | ···················· | ···················· | ···················· | ···················· | ···················· | ···················· |
| L | 3220 | hCoV-19/England/CAMC-9DFFC8/2020\|EPI_ISL_576949\|20 | ···················· | ···················· | ···················· | ···················· | ···················· | ···················· |
| L | 3221 | hCoV-19/England/CAMC-9DFFE6/2020\|EPI_ISL_576950\|20 | ···················· | ···················· | ···················· | ···················· | ···················· | ···················· |
| L | 3222 | hCoV-19/England/CAMC-9E000D/2020\|EPI_ISL_576951\|20 | ···················· | ···················· | ···················· | ···················· | ···················· | ···················· |
| L | 3223 | hCoV-19/England/CAMC-9E001C/2020\|EPI_ISL_576952\|20 | ···················· | ···················· | ···················· | ···················· | ···················· | ···················· |
| L | 3224 | hCoV-19/England/CAMC-9E003A/2020\|EPI_ISL_576953\|20 | ···················· | ···················· | ···················· | ···················· | ···················· | ···················· |
| L | 3225 | hCoV-19/England/CAMC-9E0058/2020\|EPI_ISL_576954\|20 | ···················· | ···················· | ···················· | ···················· | ···················· | ···················· |
| L | 3226 | hCoV-19/England/CAMC-9E0067/2020\|EPI_ISL_576955\|20 | ···················· | ···················· | ···················· | ···················· | ···················· | ···················· |
| L | 3227 | hCoV-19/England/CAMC-9E0085/2020\|EPI_ISL_576956\|20 | ···················· | ···················· | ···················· | ···················· | ···················· | ···················· |
| L | 3228 | hCoV-19/England/CAMC-9E0119/2020\|EPI_ISL_576958\|20 | ···················· | ···················· | ···················· | ···················· | ···················· | ···················· |
| L | 3229 | hCoV-19/England/CAMC-9E0155/2020\|EPI_ISL_576959\|20 | ···················· | ···················· | ···················· | ···················· | ···················· | ···················· |
| L | 3230 | hCoV-19/England/CAMC-9E0191/2020\|EPI_ISL_576960\|20 | ···················· | ···················· | ···················· | ···················· | ···················· | ···················· |
| L | 3231 | hCoV-19/England/LOND-D9905/2020\|EPI_ISL_576984\|202 | ···················· | ···················· | ···················· | ···················· | ···················· | ···················· |
| L | 3232 | hCoV-19/USA/LA-EVTL601/2020\|EPI_ISL_579005\|2020-06 | ···················· | ···················· | ···················· | ···················· | ···················· | ···················· |
| L | 3233 | hCoV-19/USA/LA-EVTL602/2020\|EPI_ISL_579006\|2020-06 | ···················· | ···················· | ···················· | ···················· | ···················· | ···················· |
| L | 3234 | hCoV-19/USA/LA-EVTL605/2020\|EPI_ISL_579009\|2020-08 | ···················· | ···················· | ···················· | ···················· | ···················· | ···················· |
| L | 3235 | hCoV-19/USA/LA-EVTL606/2020\|EPI_ISL_579010\|2020-07 | ···················· | ···················· | ···················· | ···················· | ···················· | ···················· |
| L | 3236 | hCoV-19/USA/LA-EVTL608/2020\|EPI_ISL_579012\|2020-08 | ···················· | ···················· | ···················· | ···················· | ···················· | ···················· |

| | | | | | | | | |
|---|---|---|---|---|---|---|---|---|
| L | 3237 | hCoV-19/USA/LA-EVTL609/2020\|EPI_ISL_579013\|2020-07 | ···················· | ···················· | ···················· | ···················· | ···················· | ···················· |
| L | 3238 | hCoV-19/USA/LA-EVTL610/2020\|EPI_ISL_579014\|2020-07 | ···················· | ···················· | ···················· | ···················· | ···················· | ···················· |
| L | 3239 | hCoV-19/USA/LA-EVTL611/2020\|EPI_ISL_579015\|2020-07 | ···················· | ···················· | ···················· | ···················· | ···················· | ···················· |
| L | 3240 | hCoV-19/USA/LA-EVTL612/2020\|EPI_ISL_579016\|2020-07 | ···················· | ···················· | ···················· | ···················· | ···················· | ···················· |
| L | 3241 | hCoV-19/USA/LA-EVTL615/2020\|EPI_ISL_579019\|2020-08 | ···················· | ···················· | ···················· | ···················· | ···················· | ···················· |
| L | 3242 | hCoV-19/New Zealand/20VR1961/2020\|EPI_ISL_579180\|2 | ···················· | ···················· | ···················· | ···················· | ···················· | ···················· |
| L | 3243 | hCoV-19/New Zealand/20VR1962/2020\|EPI_ISL_579181\|2 | ···················· | ···················· | ···················· | ···················· | ···················· | ···················· |
| L | 3245 | hCoV-19/New Zealand/20VR1964/2020\|EPI_ISL_579183\|2 | ···················· | ···················· | ···················· | ···················· | ···················· | ···················· |
| L | 3246 | hCoV-19/New Zealand/20VR1965/2020\|EPI_ISL_579184\|2 | ···················· | ···················· | ···················· | ···················· | ···················· | ···················· |
| L | 3248 | hCoV-19/New Zealand/20VR1979/2020\|EPI_ISL_579186\|2 | ···················· | ···················· | ···················· | ···················· | ···················· | ···················· |
| L | 3249 | hCoV-19/New Zealand/20VR1980/2020\|EPI_ISL_579187\|2 | ···················· | ···················· | ···················· | ···················· | ···················· | ···················· |
| L | 3250 | hCoV-19/New Zealand/20VR1981/2020\|EPI_ISL_579188\|2 | ···················· | ···················· | ···················· | ···················· | ···················· | ···················· |
| L | 3251 | hCoV-19/New Zealand/20VR1982/2020\|EPI_ISL_579189\|2 | ···················· | ···················· | ···················· | ···················· | ···················· | ···················· |
| L | 3253 | hCoV-19/New Zealand/20VR1991/2020\|EPI_ISL_579192\|2 | ···················· | ···················· | ···················· | ···················· | ···················· | ···················· |
| L | 3254 | hCoV-19/New Zealand/20VR1992/2020\|EPI_ISL_579193\|2 | ···················· | ···················· | ···················· | ···················· | ···················· | ···················· |
| L | 3255 | hCoV-19/New Zealand/20VR1993/2020\|EPI_ISL_579194\|2 | ···················· | ···················· | ···················· | ···················· | ···················· | ···················· |
| L | 3256 | hCoV-19/New Zealand/20VR1994/2020\|EPI_ISL_579195\|2 | ···················· | ···················· | ···················· | ···················· | ···················· | ···················· |
| L | 3257 | hCoV-19/New Zealand/20VR1996/2020\|EPI_ISL_579196\|2 | ···················· | ···················· | ···················· | ···················· | ···················· | ···················· |
| L | 3258 | hCoV-19/New Zealand/20VR1998/2020\|EPI_ISL_579197\|2 | ···················· | ···················· | ···················· | ···················· | ···················· | ···················· |
| L | 3259 | hCoV-19/New Zealand/20VR1999/2020\|EPI_ISL_579198\|2 | ···················· | ···················· | ···················· | ···················· | ···················· | ···················· |
| L | 3260 | hCoV-19/New Zealand/20VR2003/2020\|EPI_ISL_579199\|2 | ···················· | ···················· | ···················· | ···················· | ···················· | ···················· |
| L | 3261 | hCoV-19/England/ALDP-9E9A51/2020\|EPI_ISL_580166\|20 | ···················· | ···················· | ···················· | ···················· | ···················· | ···················· |
| L | 3262 | hCoV-19/England/ALDP-9EAF33/2020\|EPI_ISL_580167\|20 | ···················· | ···················· | ···················· | ···················· | ···················· | ···················· |
| L | 3263 | hCoV-19/England/ALDP-9EABF3/2020\|EPI_ISL_580170\|20 | ···················· | ···················· | ···················· | ···················· | ···················· | ···················· |
| L | 3264 | hCoV-19/England/ALDP-9EACB4/2020\|EPI_ISL_580171\|20 | ···················· | ···················· | ···················· | ···················· | ···················· | ···················· |
| L | 3265 | hCoV-19/England/ALDP-9E9EEC/2020\|EPI_ISL_580172\|20 | ···················· | ···················· | ···················· | ···················· | ···················· | ···················· |
| L | 3266 | hCoV-19/England/ALDP-9EA513/2020\|EPI_ISL_580173\|20 | ···················· | ···················· | ···················· | ···················· | ···················· | ···················· |
| L | 3267 | hCoV-19/England/ALDP-9EAA23/2020\|EPI_ISL_580174\|20 | ···················· | ···················· | ···················· | ···················· | ···················· | ···················· |
| L | 3268 | hCoV-19/England/ALDP-9EA258/2020\|EPI_ISL_580175\|20 | ···················· | ···················· | ···················· | ···················· | ···················· | ···················· |
| L | 3269 | hCoV-19/England/ALDP-9E9B03/2020\|EPI_ISL_580177\|20 | ···················· | ···················· | ···················· | ···················· | ···················· | ···················· |
| L | 3270 | hCoV-19/England/ALDP-9EA452/2020\|EPI_ISL_580178\|20 | ···················· | ···················· | ···················· | ···················· | ···················· | ···················· |
| L | 3271 | hCoV-19/England/ALDP-9E9E0A/2020\|EPI_ISL_580179\|20 | ···················· | ···················· | ···················· | ···················· | ···················· | ···················· |
| L | 3272 | hCoV-19/England/ALDP-9EAB7B/2020\|EPI_ISL_580181\|20 | ···················· | ···················· | ···················· | ···················· | ···················· | ···················· |
| L | 3273 | hCoV-19/England/ALDP-9EA3BF/2020\|EPI_ISL_580182\|20 | ···················· | ···················· | ···················· | ···················· | ····T··············· | ···················· |
| L | 3274 | hCoV-19/England/ALDP-9E7613/2020\|EPI_ISL_580183\|20 | ···················· | ·A·················· | ···················· | ···················· | ···················· | ···A················ |
| L | 3275 | hCoV-19/England/ALDP-9EACF0/2020\|EPI_ISL_580184\|20 | ···················· | ···················· | ···················· | ···················· | ···················· | ···················· |
| L | 3276 | hCoV-19/England/ALDP-9E9D2B/2020\|EPI_ISL_580185\|20 | ···················· | ···················· | ···················· | ···················· | ···················· | ···················· |
| L | 3277 | hCoV-19/England/ALDP-9EA1B5/2020\|EPI_ISL_580186\|20 | ···················· | ···················· | ···················· | ···················· | ···················· | ···················· |
| L | 3278 | hCoV-19/England/ALDP-9EA9CC/2020\|EPI_ISL_580188\|20 | ···················· | ···················· | ···················· | ···················· | ···················· | ···················· |
| L | 3279 | hCoV-19/England/ALDP-9E9BE5/2020\|EPI_ISL_580189\|20 | ···················· | ···················· | ···················· | ···················· | ···················· | ···················· |
| L | 3280 | hCoV-19/England/ALDP-9EA179/2020\|EPI_ISL_580190\|20 | ···················· | ···················· | ···················· | ···················· | ···················· | ···················· |
| L | 3281 | hCoV-19/England/ALDP-9E9ABB/2020\|EPI_ISL_580191\|20 | ···················· | ···················· | ···················· | ···················· | ···················· | ···················· |
| L | 3282 | hCoV-19/England/ALDP-9EA759/2020\|EPI_ISL_580192\|20 | ···················· | ···················· | ···················· | ···················· | ···················· | ···················· |
| L | 3283 | hCoV-19/England/ALDP-9EA5C8/2020\|EPI_ISL_580193\|20 | ···················· | ···················· | ···················· | ···················· | ······T············· | ···················· |
| L | 3284 | hCoV-19/England/ALDP-9E9BB8/2020\|EPI_ISL_580194\|20 | ···················· | ···················· | ···················· | ···················· | ···················· | ···················· |
| L | 3285 | hCoV-19/England/ALDP-9EAEBE/2020\|EPI_ISL_580197\|20 | ···················· | ···················· | ···················· | ···················· | ···················· | ···················· |
| L | 3286 | hCoV-19/England/ALDP-9EA7D1/2020\|EPI_ISL_580198\|20 | ···················· | ···················· | ···················· | ···················· | ···················· | ···················· |
| L | 3287 | hCoV-19/England/ALDP-9EA59B/2020\|EPI_ISL_580199\|20 | ···················· | ···················· | ···················· | ···················· | ···················· | ···················· |
| L | 3288 | hCoV-19/England/MILK-9E1F2D/2020\|EPI_ISL_581304\|20 | ···················· | ·A·················· | ···················· | ···················· | ···················· | ···A················ |
| L | 3289 | hCoV-19/England/QEUH-9DEF23/2020\|EPI_ISL_581310\|20 | ···················· | ···················· | ···················· | ···················· | ···················· | ···················· |
| L | 3290 | hCoV-19/Wales/MILK-9E329B/2020\|EPI_ISL_581311\|2020 | ···················· | ···················· | ···················· | ···················· | ···················· | ···················· |
| L | 3291 | hCoV-19/England/MILK-9E1E11/2020\|EPI_ISL_581312\|20 | ···················· | ···················· | ···················· | ···················· | ···················· | ···················· |
| L | 3292 | hCoV-19/England/MILK-9E1DBA/2020\|EPI_ISL_581313\|20 | ···················· | ···················· | ···················· | ···················· | ···················· | ···················· |
| L | 3293 | hCoV-19/Wales/MILK-9E2DB9/2020\|EPI_ISL_581314\|2020 | ···················· | ···················· | ···················· | ···················· | ···················· | ···················· |
| L | 3294 | hCoV-19/Wales/MILK-9E2AB2/2020\|EPI_ISL_581315\|2020 | ···················· | ···················· | ···················· | ···················· | ···················· | ···················· |
| L | 3295 | hCoV-19/England/MILK-9E1D50/2020\|EPI_ISL_581316\|20 | ···················· | ···················· | ···················· | ···················· | ···················· | ···················· |
| L | 3296 | hCoV-19/England/MILK-9E1F96/2020\|EPI_ISL_581317\|20 | ···················· | ···················· | ···················· | ···················· | ···················· | ···················· |
| L | 3297 | hCoV-19/England/MILK-9E1FA5/2020\|EPI_ISL_581318\|20 | ···················· | ···················· | ···················· | ···················· | ···················· | ···················· |
| L | 3298 | hCoV-19/England/MILK-9E3082/2020\|EPI_ISL_581319\|20 | ···················· | ···················· | ···················· | ···················· | ···················· | ···················· |
| L | 3299 | hCoV-19/England/MILK-9E1FC3/2020\|EPI_ISL_581320\|20 | ···················· | ···················· | ···················· | ···················· | ···················· | ···················· |
| L | 3300 | hCoV-19/England/MILK-9E201A/2020\|EPI_ISL_581321\|20 | ···················· | ···················· | ···················· | ···················· | ···················· | ···················· |
| L | 3301 | hCoV-19/England/MILK-9E21CC/2020\|EPI_ISL_581322\|20 | ···················· | ···················· | ···················· | ···················· | ···················· | ···················· |
| L | 3302 | hCoV-19/England/QEUH-9DE61F/2020\|EPI_ISL_581324\|20 | ···················· | ·A·················· | ···················· | ···················· | ···················· | ···A················ |
| L | 3303 | hCoV-19/Wales/MILK-9E2AEF/2020\|EPI_ISL_581325\|2020 | ···················· | ···················· | ···················· | ···················· | ···················· | ···················· |
| L | 3304 | hCoV-19/Wales/MILK-9E3222/2020\|EPI_ISL_581326\|2020 | ···················· | ···················· | ···················· | ···················· | ···················· | ···················· |
| L | 3305 | hCoV-19/England/MILK-9E2B82/2020\|EPI_ISL_581327\|20 | ···················· | ···················· | ···················· | ···················· | ···················· | ···················· |
| L | 3306 | hCoV-19/Wales/MILK-9E300A/2020\|EPI_ISL_581328\|2020 | ···················· | ···················· | ···················· | ···················· | ···················· | ···················· |
| L | 3307 | hCoV-19/England/QEUH-9DE767/2020\|EPI_ISL_581329\|20 | ···················· | ···················· | ···················· | ···················· | ···················· | ···················· |
| L | 3308 | hCoV-19/India/WB-IICB-021/2020\|EPI_ISL_581495\|2020 | ···················· | ···················· | ···················· | ···················· | ···················· | ···················· |
| L | 3309 | hCoV-19/India/WB-IICB-022/2020\|EPI_ISL_581496\|2020 | ···················· | ···················· | ···················· | ···················· | ···················· | ···················· |
| L | 3310 | hCoV-19/India/WB-IICB-023/2020\|EPI_ISL_581497\|2020 | ···················· | ···················· | ···················· | ···················· | ···················· | ···················· |
| L | 3311 | hCoV-19/India/WB-IICB-024/2020\|EPI_ISL_581499\|2020 | ···················· | ···················· | ···················· | ···················· | ···················· | ···················· |
| L | 3312 | hCoV-19/United Arab Emirates/skmc-0532497/2020\|EPI | ···················· | ···················· | ···················· | ···················· | ···················· | ···················· |
| L | 3313 | hCoV-19/United Arab Emirates/skmc-001/2020\|EPI_ISL | ···················· | ···················· | ···················· | ···················· | ···················· | ···················· |
| L | 3314 | hCoV-19/United Arab Emirates/skmc-003/2020\|EPI_ISL | ···················· | ···················· | ···················· | ···················· | ···················· | ···················· |
| L | 3315 | hCoV-19/United Arab Emirates/skmc-3119719/2020\|EPI | ···················· | ···················· | ···················· | ···················· | ···················· | ···················· |
| L | 3316 | hCoV-19/United Arab Emirates/skmc-3079344/2020\|EPI | ···················· | ···················· | ···················· | ···················· | ···················· | ···················· |
| L | 3317 | hCoV-19/United Arab Emirates/skmc-3111627/2020\|EPI | ···················· | ···················· | ···················· | ···················· | ···················· | ···················· |
| L | 3318 | hCoV-19/United Arab Emirates/skmc-3115511/2020\|EPI | ···················· | ···················· | ···················· | ···················· | ···················· | ···················· |
| L | 3319 | hCoV-19/United Arab Emirates/skmc-203119900/2020\|E | ···················· | ···················· | ···················· | ···················· | ···················· | ···················· |

```
L 3320 hCoV-19/United Arab Emirates/skmc-3111681/2020|EPI .................... .................... .................... .................... .................... ....................
L 3321 hCoV-19/United Arab Emirates/skmc-3125307/2020|EPI .................... .........A.......... .................... .................... .................... ....................
L 3322 hCoV-19/United Arab Emirates/skmc-3122376/2020|EPI .................... .................... .................... .................... .................... ....................
L 3323 hCoV-19/United Arab Emirates/skmc-3131248/2020|EPI .................... .................... .................... .................... .................... ....................
L 3324 hCoV-19/United Arab Emirates/skmc-203132246/2020|E .................... .................... .................... .................... .................... ....................
L 3325 hCoV-19/United Arab Emirates/skmc-3131941/2020|EPI .................... .................... .................... .................... .................... ....................
L 3326 hCoV-19/United Arab Emirates/skmc-3138700/2020|EPI .................... .A.................. .................... .................... .................... ...A................
L 3327 hCoV-19/United Arab Emirates/skmc-3124949/2020|EPI .................... .................... .................... .................... .................... ....................
L 3328 hCoV-19/United Arab Emirates/skmc-3358771/2020|EPI .................... .................... .................... .................... .................... ....................
L 3329 hCoV-19/USA/TN-QDX-1063/2020|EPI_ISL_571900|2020-0 .................... .................... .................... .................... .................... ....................
L 3330 hCoV-19/USA/TX-QDX-1062/2020|EPI_ISL_571901|2020-0 .................... .................... .................... .................... .................... ....................
L 3331 hCoV-19/USA/TX-QDX-1096/2020|EPI_ISL_571902|2020-0 .................... .................... .................... .................... .................... ....................
L 3332 hCoV-19/USA/TX-QDX-1097/2020|EPI_ISL_571903|2020-0 .................... .................... .................... .................... .................... ....................
L 3333 hCoV-19/USA/WA-QDX-1080/2020|EPI_ISL_571905|2020-0 .................... .................... .................... .................... .................... ....................
L 3334 hCoV-19/USA/WA-QDX-1084/2020|EPI_ISL_571906|2020-0 .................... .................... .................... .................... .................... ....................
L 3335 hCoV-19/USA/WA-QDX-1085/2020|EPI_ISL_571907|2020-0 .................... .................... .................... .................... .................... ....................
L 3336 hCoV-19/USA/WA-QDX-1104/2020|EPI_ISL_571908|2020-0 .................... .................... .................... .................... .................... ....................
L 3337 hCoV-19/USA/WA-QDX-1118/2020|EPI_ISL_571909|2020-0 .................... .................... .................... .................... .................... ....................
L 3338 hCoV-19/USA/WI-QDX-1072/2020|EPI_ISL_571910|2020-0 .................... .................... .................... .................... .................... ....................
L 3339 hCoV-19/USA/WI-QDX-1076/2020|EPI_ISL_571912|2020-0 .................... .................... .................... .................... .................... ....................
L 3340 hCoV-19/USA/WI-QDX-1077/2020|EPI_ISL_571913|2020-0 .................... .................... .................... .................... .................... ....................
L 3341 hCoV-19/USA/WY-QDX-1066/2020|EPI_ISL_571915|2020-0 .................... .................... .................... .................... .................... ....................
L 3342 hCoV-19/USA/AR-QDX-1249/2020|EPI_ISL_571916|2020-0 .................... .................... .................... .................... .................... ....................
L 3343 hCoV-19/USA/CA-QDX-1244/2020|EPI_ISL_571917|2020-0 .................... .................... .................... .................... .................... ....................
L 3344 hCoV-19/USA/CA-QDX-1245/2020|EPI_ISL_571918|2020-0 .................... .................... .................... .................... .................... ....................
L 3345 hCoV-19/USA/CA-QDX-1246/2020|EPI_ISL_571919|2020-0 .................... .................... .................... .................... .................... ....................
L 3346 hCoV-19/USA/CA-QDX-1247/2020|EPI_ISL_571920|2020-0 .................... .................... .................... .................... .................... ....................
L 3347 hCoV-19/USA/CA-QDX-1248/2020|EPI_ISL_571921|2020-0 .................... .................... .................... .................... .................... ....................
L 3348 hCoV-19/USA/CA-QDX-1286/2020|EPI_ISL_571922|2020-0 .................... .................... .................... .................... .................... ....................
L 3349 hCoV-19/USA/CA-QDX-1287/2020|EPI_ISL_571923|2020-0 .................... .................... .................... .................... .................... ....................
L 3350 hCoV-19/USA/CA-QDX-1289/2020|EPI_ISL_571924|2020-0 .................... .................... .................... .................... .................... ....................
L 3351 hCoV-19/USA/CO-QDX-1312/2020|EPI_ISL_571925|2020-0 .................... ..........A......... .................... .................... .................... ....................
L 3352 hCoV-19/USA/CT-QDX-1183/2020|EPI_ISL_571926|2020-0 .................... .................... .................... .................... .................... ....................
L 3353 hCoV-19/USA/ID-QDX-1272/2020|EPI_ISL_571928|2020-0 .................... .................... .................... .................... .................... ....................
L 3354 hCoV-19/USA/ID-QDX-1290/2020|EPI_ISL_571929|2020-0 .................... .................... .................... .................... .................... ....................
L 3355 hCoV-19/USA/ID-QDX-1291/2020|EPI_ISL_571930|2020-0 .................... .................... .................... .................... .................... ....................
L 3356 hCoV-19/USA/ID-QDX-1292/2020|EPI_ISL_571931|2020-0 .................... .................... .................... .................... .................... ....................
L 3357 hCoV-19/USA/ID-QDX-1293/2020|EPI_ISL_571932|2020-0 .................... .................... .................... .................... .................... ....................
L 3358 hCoV-19/USA/ID-QDX-1294/2020|EPI_ISL_571933|2020-0 .................... .................... .................... .................... .................... ....................
L 3359 hCoV-19/USA/IN-QDX-1186/2020|EPI_ISL_571934|2020-0 .................... .................... .................... .................... .................... ....................
L 3360 hCoV-19/USA/IN-QDX-1188/2020|EPI_ISL_571936|2020-0 .................... .................... .................... .................... .................... ....................
L 3361 hCoV-19/USA/IN-QDX-1189/2020|EPI_ISL_571937|2020-0 .................... .................... .................... .................... .................... ....................
L 3362 hCoV-19/USA/LA-QDX-1192/2020|EPI_ISL_571938|2020-0 .................... .................... .................... .................... .................... ....................
L 3363 hCoV-19/USA/LA-QDX-1193/2020|EPI_ISL_571939|2020-0 .................... .................... .................... .................... .................... ....................
L 3364 hCoV-19/USA/LA-QDX-1194/2020|EPI_ISL_571940|2020-0 .................... .................... .................... .................... .................... ....................
L 3365 hCoV-19/USA/LA-QDX-1195/2020|EPI_ISL_571941|2020-0 .................... .................... .................... .................... .................... ....................
L 3366 hCoV-19/USA/LA-QDX-1196/2020|EPI_ISL_571942|2020-0 .................... .................... .................... .................... .................... ....................
L 3367 hCoV-19/USA/LA-QDX-1200/2020|EPI_ISL_571943|2020-0 .................... .................... .................... .................... .................... ....................
L 3368 hCoV-19/USA/LA-QDX-1203/2020|EPI_ISL_571944|2020-0 .................... .................... .................... .................... .................... ....................
L 3369 hCoV-19/USA/LA-QDX-1204/2020|EPI_ISL_571945|2020-0 .................... .................... .................... .................... .................... ....................
L 3370 hCoV-19/USA/LA-QDX-1205/2020|EPI_ISL_571946|2020-0 .................... .................... .................... .................... .................... ....................
L 3371 hCoV-19/USA/LA-QDX-1206/2020|EPI_ISL_571947|2020-0 .................... .................... .................... .................... .................... ....................
L 3372 hCoV-19/USA/LA-QDX-1241/2020|EPI_ISL_571949|2020-0 .................... .................... .................... .................... .................... ....................
L 3373 hCoV-19/USA/LA-QDX-1242/2020|EPI_ISL_571950|2020-0 .................... .................... .................... .................... .................... ....................
L 3374 hCoV-19/USA/MI-QDX-1295/2020|EPI_ISL_571951|2020-0 .................... .................... .................... .................... .................... ....................
L 3375 hCoV-19/USA/MI-QDX-1296/2020|EPI_ISL_571952|2020-0 .................... .................... .................... .................... .................... ....................
L 3376 hCoV-19/USA/MI-QDX-1297/2020|EPI_ISL_571953|2020-0 .................... .................... .................... .................... .................... ....................
L 3377 hCoV-19/USA/MI-QDX-1299/2020|EPI_ISL_571954|2020-0 .................... .................... .................... .................... .................... ....................
L 3378 hCoV-19/USA/MI-QDX-1300/2020|EPI_ISL_571955|2020-0 .................... .................... .................... .................... .................... ....................
L 3379 hCoV-19/USA/MI-QDX-1301/2020|EPI_ISL_571956|2020-0 .................... .................... .................... .................... .................... ....................
L 3380 hCoV-19/USA/MI-QDX-1302/2020|EPI_ISL_571957|2020-0 .................... .................... .................... .................... .................... ....................
L 3381 hCoV-19/USA/MI-QDX-1303/2020|EPI_ISL_571958|2020-0 .................... .................... .................... .................... .................... ....................
L 3382 hCoV-19/USA/MI-QDX-1304/2020|EPI_ISL_571959|2020-0 .................... .................... .................... .................... .................... ....................
L 3383 hCoV-19/USA/MI-QDX-1305/2020|EPI_ISL_571960|2020-0 .................... .................... .................... .................... .................... ....................
L 3384 hCoV-19/USA/MI-QDX-1306/2020|EPI_ISL_571961|2020-0 .................... .................... .................... .................... .................... ....................
L 3385 hCoV-19/USA/MI-QDX-1307/2020|EPI_ISL_571962|2020-0 .................... .................... .................... .................... .................... ....................
L 3386 hCoV-19/USA/MI-QDX-1308/2020|EPI_ISL_571963|2020-0 .................... .................... .................... .................... .................... ....................
L 3387 hCoV-19/USA/MI-QDX-1309/2020|EPI_ISL_571964|2020-0 .................... .................... .................... .................... .................... ....................
L 3388 hCoV-19/USA/MI-QDX-1310/2020|EPI_ISL_571965|2020-0 .................... .................... .................... .................... .................... ....................
L 3389 hCoV-19/USA/MI-QDX-1311/2020|EPI_ISL_571966|2020-0 .................... .................... .................... .................... .................... ....................
L 3390 hCoV-19/USA/MI-QDX-1313/2020|EPI_ISL_571967|2020-0 .................... .................... .................... .................... .................... ....................
L 3391 hCoV-19/USA/MS-QDX-1243/2020|EPI_ISL_571968|2020-0 .................... .................... .................... .................... .................... ....................
L 3392 hCoV-19/USA/NJ-QDX-1185/2020|EPI_ISL_571969|2020-0 .................... .................... .................... .................... .................... ....................
L 3393 hCoV-19/USA/NY-QDX-1181/2020|EPI_ISL_571991|2020-0 .................... .................... .................... .................... .................... ....................
L 3394 hCoV-19/USA/NY-QDX-1182/2020|EPI_ISL_571992|2020-0 .................... .................... .................... .................... .................... ....................
L 3395 hCoV-19/USA/NY-QDX-1184/2020|EPI_ISL_571993|2020-0 .................... .................... .................... .................... .................... ....................
L 3396 hCoV-19/USA/NY-QDX-1214/2020|EPI_ISL_571994|2020-0 .................... .................... .................... .................... .................... ....................
L 3397 hCoV-19/USA/NY-QDX-1215/2020|EPI_ISL_571995|2020-0 .................... .................... .................... .................... .................... ....................
L 3398 hCoV-19/USA/NY-QDX-1218/2020|EPI_ISL_571996|2020-0 .................... .................... .................... .................... .................... ....................
L 3399 hCoV-19/USA/NY-QDX-1219/2020|EPI_ISL_571997|2020-0 .................... .................... .................... .................... .................... ....................
```

<table>
<tr><td>L</td><td>3400</td><td>hCoV-19/USA/NY-QDX-1220/2020|EPI_ISL_571998|2020-0</td><td>····················</td><td>····················</td><td>····················</td><td>····················</td><td>····················</td><td>····················</td><td></td></tr>
<tr><td>L</td><td>3401</td><td>hCoV-19/USA/NY-QDX-1222/2020|EPI_ISL_571999|2020-0</td><td>····················</td><td>···················A</td><td>····················</td><td>····················</td><td>····················</td><td>····················</td><td>1</td></tr>
<tr><td>L</td><td>3402</td><td>hCoV-19/Wales/PHWC-16CDDD/2020|EPI_ISL_574169|2020</td><td>····················</td><td>····················</td><td>····················</td><td>····················</td><td>····················</td><td>····················</td><td></td></tr>
<tr><td>L</td><td>3403</td><td>hCoV-19/Wales/PHWC-16CDEC/2020|EPI_ISL_574170|2020</td><td>····················</td><td>····················</td><td>····················</td><td>····················</td><td>····················</td><td>····················</td><td></td></tr>
<tr><td>L</td><td>3404</td><td>hCoV-19/Wales/PHWC-16CDFB/2020|EPI_ISL_574171|2020</td><td>····················</td><td>····················</td><td>····················</td><td>····················</td><td>····················</td><td>····················</td><td></td></tr>
<tr><td>L</td><td>3405</td><td>hCoV-19/Wales/PHWC-16CE07/2020|EPI_ISL_574172|2020</td><td>····················</td><td>····················</td><td>····················</td><td>····················</td><td>····················</td><td>····················</td><td></td></tr>
<tr><td>L</td><td>3406</td><td>hCoV-19/Wales/PHWC-16CE16/2020|EPI_ISL_574173|2020</td><td>····················</td><td>····················</td><td>····················</td><td>····················</td><td>····················</td><td>····················</td><td></td></tr>
<tr><td>L</td><td>3407</td><td>hCoV-19/Wales/PHWC-16CE34/2020|EPI_ISL_574174|2020</td><td>····················</td><td>····················</td><td>····················</td><td>····················</td><td>····················</td><td>····················</td><td></td></tr>
<tr><td>L</td><td>3408</td><td>hCoV-19/Wales/PHWC-16CE43/2020|EPI_ISL_574175|2020</td><td>····················</td><td>····················</td><td>····················</td><td>····················</td><td>····················</td><td>····················</td><td></td></tr>
<tr><td>L</td><td>3409</td><td>hCoV-19/Wales/PHWC-16CE52/2020|EPI_ISL_574176|2020</td><td>····················</td><td>····················</td><td>····················</td><td>····················</td><td>····················</td><td>····················</td><td></td></tr>
<tr><td>L</td><td>3410</td><td>hCoV-19/Wales/PHWC-16CE70/2020|EPI_ISL_574177|2020</td><td>····················</td><td>····················</td><td>····················</td><td>····················</td><td>····················</td><td>····················</td><td></td></tr>
<tr><td>L</td><td>3411</td><td>hCoV-19/Wales/PHWC-16CE8F/2020|EPI_ISL_574178|2020</td><td>····················</td><td>····················</td><td>····················</td><td>····················</td><td>····················</td><td>····················</td><td></td></tr>
<tr><td>L</td><td>3412</td><td>hCoV-19/Wales/PHWC-16CE9E/2020|EPI_ISL_574179|2020</td><td>····················</td><td>····················</td><td>····················</td><td>····················</td><td>····················</td><td>····················</td><td></td></tr>
<tr><td>L</td><td>3413</td><td>hCoV-19/Wales/PHWC-16CEBC/2020|EPI_ISL_574180|2020</td><td>····················</td><td>····················</td><td>····················</td><td>····················</td><td>····················</td><td>····················</td><td></td></tr>
<tr><td>L</td><td>3417</td><td>hCoV-19/Wales/PHWC-16CF04/2020|EPI_ISL_574184|2020</td><td>····················</td><td>····················</td><td>····················</td><td>····················</td><td>····················</td><td>····················</td><td></td></tr>
<tr><td>L</td><td>3418</td><td>hCoV-19/Wales/PHWC-16CF13/2020|EPI_ISL_574185|2020</td><td>····················</td><td>····················</td><td>····················</td><td>····················</td><td>····················</td><td>····················</td><td></td></tr>
<tr><td>L</td><td>3419</td><td>hCoV-19/Wales/PHWC-16CF22/2020|EPI_ISL_574186|2020</td><td>····················</td><td>····················</td><td>····················</td><td>····················</td><td>····················</td><td>····················</td><td></td></tr>
<tr><td>L</td><td>3420</td><td>hCoV-19/Wales/PHWC-16CF31/2020|EPI_ISL_574187|2020</td><td>····················</td><td>····················</td><td>····················</td><td>····················</td><td>NNN·················</td><td>····················</td><td>1</td></tr>
<tr><td>L</td><td>3421</td><td>hCoV-19/Wales/PHWC-16CF5F/2020|EPI_ISL_574188|2020</td><td>····················</td><td>····················</td><td>····················</td><td>····················</td><td>····················</td><td>····················</td><td></td></tr>
<tr><td>L</td><td>3422</td><td>hCoV-19/Wales/PHWC-16CF8C/2020|EPI_ISL_574189|2020</td><td>····················</td><td>····················</td><td>····················</td><td>····················</td><td>····················</td><td>····················</td><td></td></tr>
<tr><td>L</td><td>3424</td><td>hCoV-19/Wales/PHWC-16CFAA/2020|EPI_ISL_574191|2020</td><td>····················</td><td>····················</td><td>····················</td><td>····················</td><td>····················</td><td>····················</td><td></td></tr>
<tr><td>L</td><td>3425</td><td>hCoV-19/Wales/PHWC-16CFB9/2020|EPI_ISL_574192|2020</td><td>····················</td><td>····················</td><td>····················</td><td>····················</td><td>····················</td><td>····················</td><td></td></tr>
<tr><td>L</td><td>3426</td><td>hCoV-19/Wales/PHWC-16CFD7/2020|EPI_ISL_574193|2020</td><td>····················</td><td>····················</td><td>····················</td><td>····················</td><td>····················</td><td>····················</td><td></td></tr>
<tr><td>L</td><td>3427</td><td>hCoV-19/Wales/PHWC-16CFE6/2020|EPI_ISL_574194|2020</td><td>····················</td><td>····················</td><td>····················</td><td>····················</td><td>····················</td><td>····················</td><td></td></tr>
<tr><td>L</td><td>3429</td><td>hCoV-19/Wales/PHWC-16D000/2020|EPI_ISL_574196|2020</td><td>····················</td><td>····················</td><td>····················</td><td>····················</td><td>····················</td><td>····················</td><td></td></tr>
<tr><td>L</td><td>3430</td><td>hCoV-19/Wales/PHWC-16D04C/2020|EPI_ISL_574197|2020</td><td>····················</td><td>····················</td><td>····················</td><td>····················</td><td>····················</td><td>····················</td><td></td></tr>
<tr><td>L</td><td>3431</td><td>hCoV-19/Wales/PHWC-16D05B/2020|EPI_ISL_574198|2020</td><td>····················</td><td>····················</td><td>····················</td><td>····················</td><td>····················</td><td>····················</td><td></td></tr>
<tr><td>L</td><td>3432</td><td>hCoV-19/Wales/PHWC-16D097/2020|EPI_ISL_574199|2020</td><td>····················</td><td>····················</td><td>····················</td><td>····················</td><td>····················</td><td>····················</td><td></td></tr>
<tr><td>L</td><td>3434</td><td>hCoV-19/England/ALDP-9CB958/2020|EPI_ISL_575550|20</td><td>····················</td><td>····················</td><td>····················</td><td>····················</td><td>····················</td><td>····················</td><td></td></tr>
<tr><td>L</td><td>3435</td><td>hCoV-19/England/ALDP-9CB994/2020|EPI_ISL_575554|20</td><td>····················</td><td>····················</td><td>····················</td><td>····················</td><td>····················</td><td>····················</td><td></td></tr>
<tr><td>L</td><td>3436</td><td>hCoV-19/England/BIRM-63D04/2020|EPI_ISL_576880|202</td><td>····················</td><td>····················</td><td>····················</td><td>····················</td><td>····················</td><td>····················</td><td></td></tr>
<tr><td>L</td><td>3437</td><td>hCoV-19/England/BIRM-63D31/2020|EPI_ISL_576881|202</td><td>····················</td><td>····················</td><td>····················</td><td>····················</td><td>····················</td><td>····················</td><td></td></tr>
<tr><td>L</td><td>3438</td><td>hCoV-19/England/BIRM-63DD7/2020|EPI_ISL_576885|202</td><td>····················</td><td>····················</td><td>····················</td><td>····················</td><td>····················</td><td>····················</td><td></td></tr>
<tr><td>L</td><td>3439</td><td>hCoV-19/USA/LA-EVTL596/2020|EPI_ISL_579000|2020-06</td><td>····················</td><td>····················</td><td>····················</td><td>····················</td><td>····················</td><td>····················</td><td></td></tr>
<tr><td>L</td><td>3440</td><td>hCoV-19/USA/LA-EVTL597/2020|EPI_ISL_579001|2020-06</td><td>····················</td><td>····················</td><td>····················</td><td>····················</td><td>····················</td><td>····················</td><td></td></tr>
<tr><td>L</td><td>3441</td><td>hCoV-19/USA/LA-EVTL598/2020|EPI_ISL_579002|2020-06</td><td>····················</td><td>····················</td><td>····················</td><td>····················</td><td>····················</td><td>····················</td><td></td></tr>
<tr><td>L</td><td>3442</td><td>hCoV-19/USA/LA-EVTL599/2020|EPI_ISL_579003|2020-06</td><td>····················</td><td>····················</td><td>····················</td><td>····················</td><td>····················</td><td>····················</td><td></td></tr>
<tr><td>L</td><td>3443</td><td>hCoV-19/USA/LA-EVTL600/2020|EPI_ISL_579004|2020-06</td><td>····················</td><td>····················</td><td>····················</td><td>····················</td><td>····················</td><td>····················</td><td></td></tr>
<tr><td>L</td><td>3444</td><td>hCoV-19/USA/LA-EVTL603/2020|EPI_ISL_579007|2020-06</td><td>····················</td><td>····················</td><td>····················</td><td>····················</td><td>····················</td><td>····················</td><td></td></tr>
<tr><td>L</td><td>3445</td><td>hCoV-19/USA/LA-EVTL604/2020|EPI_ISL_579008|2020-08</td><td>····················</td><td>····················</td><td>····················</td><td>····················</td><td>····················</td><td>····················</td><td></td></tr>
<tr><td>L</td><td>3446</td><td>hCoV-19/USA/LA-EVTL616/2020|EPI_ISL_579020|2020-07</td><td>····················</td><td>····················</td><td>····················</td><td>····················</td><td>····················</td><td>····················</td><td></td></tr>
<tr><td>L</td><td>3447</td><td>hCoV-19/USA/LA-EVTL617/2020|EPI_ISL_579021|2020-08</td><td>····················</td><td>····················</td><td>····················</td><td>····················</td><td>····················</td><td>····················</td><td></td></tr>
<tr><td>L</td><td>3448</td><td>hCoV-19/USA/LA-EVTL618/2020|EPI_ISL_579022|2020-07</td><td>····················</td><td>····················</td><td>····················</td><td>····················</td><td>····················</td><td>····················</td><td></td></tr>
<tr><td>L</td><td>3449</td><td>hCoV-19/USA/LA-EVTL619/2020|EPI_ISL_579023|2020-08</td><td>····················</td><td>····················</td><td>····················</td><td>····················</td><td>····················</td><td>····················</td><td></td></tr>
<tr><td>L</td><td>3450</td><td>hCoV-19/USA/LA-EVTL620/2020|EPI_ISL_579024|2020-07</td><td>····················</td><td>····················</td><td>····················</td><td>····················</td><td>····················</td><td>····················</td><td></td></tr>
<tr><td>L</td><td>3451</td><td>hCoV-19/USA/LA-EVTL621/2020|EPI_ISL_579025|2020-07</td><td>····················</td><td>····················</td><td>····················</td><td>····················</td><td>····················</td><td>····················</td><td></td></tr>
<tr><td>L</td><td>3452</td><td>hCoV-19/USA/LA-EVTL622/2020|EPI_ISL_579026|2020-08</td><td>····················</td><td>····················</td><td>····················</td><td>····················</td><td>····················</td><td>····················</td><td></td></tr>
<tr><td>L</td><td>3453</td><td>hCoV-19/USA/LA-EVTL623/2020|EPI_ISL_579027|2020-07</td><td>····················</td><td>····················</td><td>····················</td><td>····················</td><td>····················</td><td>····················</td><td></td></tr>
<tr><td>L</td><td>3454</td><td>hCoV-19/USA/LA-EVTL624/2020|EPI_ISL_579028|2020-07</td><td>····················</td><td>····················</td><td>····················</td><td>····················</td><td>····················</td><td>····················</td><td></td></tr>
<tr><td>L</td><td>3455</td><td>hCoV-19/USA/LA-EVTL625/2020|EPI_ISL_579029|2020-08</td><td>····················</td><td>····················</td><td>····················</td><td>····················</td><td>····················</td><td>····················</td><td></td></tr>
<tr><td>L</td><td>3456</td><td>hCoV-19/USA/LA-EVTL626/2020|EPI_ISL_579030|2020-07</td><td>····················</td><td>····················</td><td>····················</td><td>····················</td><td>····················</td><td>····················</td><td></td></tr>
<tr><td>L</td><td>3457</td><td>hCoV-19/USA/LA-EVTL627/2020|EPI_ISL_579031|2020-07</td><td>····················</td><td>····················</td><td>····················</td><td>····················</td><td>····················</td><td>····················</td><td></td></tr>
<tr><td>L</td><td>3458</td><td>hCoV-19/USA/LA-EVTL628/2020|EPI_ISL_579032|2020-08</td><td>····················</td><td>····················</td><td>····················</td><td>····················</td><td>····················</td><td>····················</td><td></td></tr>
<tr><td>L</td><td>3459</td><td>hCoV-19/USA/LA-EVTL629/2020|EPI_ISL_579033|2020-08</td><td>····················</td><td>····················</td><td>····················</td><td>····················</td><td>····················</td><td>····················</td><td></td></tr>
<tr><td>L</td><td>3460</td><td>hCoV-19/USA/LA-EVTL630/2020|EPI_ISL_579034|2020-07</td><td>····················</td><td>····················</td><td>····················</td><td>····················</td><td>····················</td><td>····················</td><td></td></tr>
<tr><td>L</td><td>3461</td><td>hCoV-19/USA/LA-EVTL631/2020|EPI_ISL_579035|2020-08</td><td>····················</td><td>····················</td><td>····················</td><td>····················</td><td>····················</td><td>····················</td><td></td></tr>
<tr><td>L</td><td>3462</td><td>hCoV-19/USA/LA-EVTL632/2020|EPI_ISL_579036|2020-08</td><td>····················</td><td>······A·············</td><td>····················</td><td>····················</td><td>····················</td><td>····················</td><td></td></tr>
<tr><td>L</td><td>3463</td><td>hCoV-19/USA/LA-EVTL633/2020|EPI_ISL_579037|2020-08</td><td>····················</td><td>······A·············</td><td>····················</td><td>····················</td><td>····················</td><td>····················</td><td></td></tr>
<tr><td>L</td><td>3464</td><td>hCoV-19/USA/LA-EVTL634/2020|EPI_ISL_579038|2020-08</td><td>····················</td><td>····················</td><td>····················</td><td>····················</td><td>····················</td><td>····················</td><td></td></tr>
<tr><td>L</td><td>3465</td><td>hCoV-19/USA/LA-EVTL635/2020|EPI_ISL_579039|2020-08</td><td>····················</td><td>······A·············</td><td>····················</td><td>····················</td><td>····················</td><td>····················</td><td></td></tr>
<tr><td>L</td><td>3466</td><td>hCoV-19/USA/LA-EVTL636/2020|EPI_ISL_579040|2020-08</td><td>····················</td><td>····················</td><td>····················</td><td>····················</td><td>····················</td><td>····················</td><td></td></tr>
<tr><td>L</td><td>3467</td><td>hCoV-19/USA/LA-EVTL637/2020|EPI_ISL_579041|2020-07</td><td>····················</td><td>····················</td><td>····················</td><td>····················</td><td>····················</td><td>····················</td><td></td></tr>
<tr><td>L</td><td>3468</td><td>hCoV-19/USA/LA-EVTL638/2020|EPI_ISL_579042|2020-08</td><td>····················</td><td>····················</td><td>····················</td><td>····················</td><td>····················</td><td>····················</td><td></td></tr>
<tr><td>L</td><td>3469</td><td>hCoV-19/USA/LA-EVTL639/2020|EPI_ISL_579043|2020-08</td><td>····················</td><td>····················</td><td>····················</td><td>····················</td><td>····················</td><td>····················</td><td></td></tr>
<tr><td>L</td><td>3470</td><td>hCoV-19/USA/LA-EVTL640/2020|EPI_ISL_579044|2020-08</td><td>····················</td><td>····················</td><td>····················</td><td>····················</td><td>····················</td><td>····················</td><td></td></tr>
<tr><td>L</td><td>3471</td><td>hCoV-19/USA/LA-EVTL641/2020|EPI_ISL_579045|2020-08</td><td>····················</td><td>····················</td><td>····················</td><td>····················</td><td>····················</td><td>····················</td><td></td></tr>
<tr><td>L</td><td>3472</td><td>hCoV-19/USA/LA-EVTL642/2020|EPI_ISL_579046|2020-08</td><td>····················</td><td>····················</td><td>····················</td><td>····················</td><td>····················</td><td>····················</td><td></td></tr>
<tr><td>L</td><td>3473</td><td>hCoV-19/USA/LA-EVTL643/2020|EPI_ISL_579047|2020-08</td><td>····················</td><td>······A·············</td><td>····················</td><td>····················</td><td>····················</td><td>····················</td><td></td></tr>
<tr><td>L</td><td>3474</td><td>hCoV-19/USA/LA-EVTL644/2020|EPI_ISL_579048|2020-08</td><td>····················</td><td>······A·············</td><td>····················</td><td>····················</td><td>····················</td><td>····················</td><td></td></tr>
<tr><td>L</td><td>3475</td><td>hCoV-19/USA/LA-EVTL645/2020|EPI_ISL_579049|2020-08</td><td>····················</td><td>······A·············</td><td>····················</td><td>····················</td><td>····················</td><td>····················</td><td></td></tr>
<tr><td>L</td><td>3476</td><td>hCoV-19/USA/LA-EVTL646/2020|EPI_ISL_579050|2020-08</td><td>····················</td><td>····················</td><td>····················</td><td>····················</td><td>····················</td><td>····················</td><td></td></tr>
<tr><td>L</td><td>3477</td><td>hCoV-19/USA/LA-EVTL647/2020|EPI_ISL_579051|2020-08</td><td>····················</td><td>····················</td><td>····················</td><td>····················</td><td>····················</td><td>····················</td><td></td></tr>
<tr><td>L</td><td>3478</td><td>hCoV-19/USA/LA-EVTL648/2020|EPI_ISL_579052|2020-08</td><td>····················</td><td>····················</td><td>····················</td><td>····················</td><td>····················</td><td>····················</td><td></td></tr>
<tr><td>L</td><td>3479</td><td>hCoV-19/USA/LA-EVTL649/2020|EPI_ISL_579053|2020-08</td><td>····················</td><td>····················</td><td>····················</td><td>····················</td><td>····················</td><td>····················</td><td></td></tr>
<tr><td>L</td><td>3480</td><td>hCoV-19/USA/LA-EVTL650/2020|EPI_ISL_579054|2020-08</td><td>····················</td><td>····················</td><td>····················</td><td>····················</td><td>····················</td><td>····················</td><td></td></tr>
<tr><td>L</td><td>3481</td><td>hCoV-19/USA/LA-EVTL651/2020|EPI_ISL_579055|2020-08</td><td>····················</td><td>····················</td><td>····················</td><td>····················</td><td>····················</td><td>····················</td><td></td></tr>
<tr><td>L</td><td>3482</td><td>hCoV-19/USA/LA-EVTL652/2020|EPI_ISL_579056|2020-08</td><td>····················</td><td>····················</td><td>····················</td><td>····················</td><td>····················</td><td>····················</td><td></td></tr>
<tr><td>L</td><td>3483</td><td>hCoV-19/USA/LA-EVTL185/2020|EPI_ISL_579057|2020-08</td><td>····················</td><td>····················</td><td>····················</td><td>····················</td><td>····················</td><td>····················</td><td></td></tr>
<tr><td>L</td><td>3484</td><td>hCoV-19/England/ALDP-9EAD2A/2020|EPI_ISL_580000|20</td><td>····················</td><td>····················</td><td>····················</td><td>····················</td><td>····················</td><td>····················</td><td></td></tr>
<tr><td>L</td><td>3485</td><td>hCoV-19/England/ALDP-9EABD5/2020|EPI_ISL_580001|20</td><td>····················</td><td>·A··················</td><td>····················</td><td>····················</td><td>····················</td><td>··A·················</td><td></td></tr>
</table>

| | | | | | | | | |
|---|---|---|---|---|---|---|---|---|
| L | 3486 | hCoV-19/England/ALDP-9E9A7F/2020\|EPI_ISL_580002\|20 | ···················· | ···················· | ····················· | ···················· | ···················· | ···················· |
| L | 3487 | hCoV-19/England/ALDP-9EA6F2/2020\|EPI_ISL_580003\|20 | ···················· | ···················· | ····················· | ···················· | ···················· | ···················· |
| L | 3488 | hCoV-19/England/ALDP-9EA57D/2020\|EPI_ISL_580004\|20 | ···················· | ···················· | ····················· | ···················· | ···················· | ···················· |
| L | 3489 | hCoV-19/England/ALDP-9EAA41/2020\|EPI_ISL_580006\|20 | ···················· | ···················· | ····················· | ···················· | ···················· | ···················· |
| L | 3490 | hCoV-19/England/ALDP-9E9B21/2020\|EPI_ISL_580007\|20 | ···················· | ·A·················· | ····················· | ···················· | ···················· | ···················· |
| L | 3491 | hCoV-19/England/ALDP-9EAE90/2020\|EPI_ISL_580008\|20 | ···················· | ···················· | ····················· | ···················· | ···················· | ···················· |
| L | 3492 | hCoV-19/England/ALDP-9EAF24/2020\|EPI_ISL_580009\|20 | ···················· | ···················· | ····················· | ···················· | ···················· | ···················· |
| L | 3493 | hCoV-19/England/ALDP-9EA5B9/2020\|EPI_ISL_580010\|20 | ···················· | ···················· | ····················· | ···················· | ···················· | ···················· |
| L | 3494 | hCoV-19/England/ALDP-9EA610/2020\|EPI_ISL_580011\|20 | ···················· | ···················· | ····················· | ···················· | ···················· | ···················· |
| L | 3495 | hCoV-19/England/ALDP-9EAF15/2020\|EPI_ISL_580012\|20 | ···················· | ···················· | ····················· | ···················· | ···················· | ···················· |
| L | 3496 | hCoV-19/England/ALDP-9EA003/2020\|EPI_ISL_580013\|20 | ···················· | ···················· | ····················· | ···················· | ···················· | ···················· |
| L | 3497 | hCoV-19/England/ALDP-9E9C97/2020\|EPI_ISL_580014\|20 | ···················· | ···················· | ····················· | ···················· | ···················· | ···················· |
| L | 3498 | hCoV-19/England/ALDP-9EAFCA/2020\|EPI_ISL_580016\|20 | ···················· | ···················· | ····················· | ···················· | ···················· | ···················· |
| L | 3499 | hCoV-19/England/ALDP-9EA971/2020\|EPI_ISL_580017\|20 | ···················· | ···················· | ····················· | ···················· | ···················· | ···················· |
| L | 3500 | hCoV-19/England/ALDP-9EA1A6/2020\|EPI_ISL_580018\|20 | ···················· | ···················· | ····················· | ···················· | ···················· | ···················· |
| L | 3501 | hCoV-19/England/ALDP-9EA425/2020\|EPI_ISL_580019\|20 | ···················· | ···················· | ····················· | ···················· | ···················· | ···················· |
| L | 3502 | hCoV-19/England/ALDP-9EA74A/2020\|EPI_ISL_580020\|20 | ···················· | ···················· | ····················· | ···················· | ···················· | ···················· |
| L | 3503 | hCoV-19/England/ALDP-9EAE81/2020\|EPI_ISL_580021\|20 | ···················· | ···················· | ····················· | ···················· | ···················· | ···················· |
| L | 3504 | hCoV-19/England/ALDP-9E9BF4/2020\|EPI_ISL_580022\|20 | ···················· | ···················· | ····················· | ···················· | ···················· | ···················· |
| L | 3505 | hCoV-19/England/ALDP-9EABA8/2020\|EPI_ISL_580023\|20 | ···················· | ···················· | ····················· | ···················· | ···················· | ···················· |
| L | 3506 | hCoV-19/England/ALDP-9EA944/2020\|EPI_ISL_580024\|20 | ···················· | ···················· | ····················· | ···················· | ···················· | ···················· |
| L | 3507 | hCoV-19/England/ALDP-9EA65C/2020\|EPI_ISL_580025\|20 | ···················· | ···················· | ····················· | ···················· | ···················· | ···················· |
| L | 3508 | hCoV-19/England/ALDP-9EAB5D/2020\|EPI_ISL_580026\|20 | ···················· | ···················· | ····················· | ···················· | ···················· | ···················· |
| L | 3509 | hCoV-19/England/ALDP-9EA9EA/2020\|EPI_ISL_580028\|20 | ···················· | ···················· | ····················· | ···················· | ···················· | ···················· |
| L | 3510 | hCoV-19/England/ALDP-9EAE54/2020\|EPI_ISL_580029\|20 | ···················· | ···················· | ····················· | ···················· | ···················· | ···················· |
| L | 3511 | hCoV-19/England/ALDP-9EA2B2/2020\|EPI_ISL_580030\|20 | ···················· | ···················· | ····················· | ···················· | ···················· | ···················· |
| L | 3512 | hCoV-19/England/ALDP-9EA2D0/2020\|EPI_ISL_580031\|20 | ···················· | ···················· | ····················· | ···················· | ···················· | ···················· |
| L | 3513 | hCoV-19/England/ALDP-9E9EDD/2020\|EPI_ISL_580032\|20 | ···················· | ···················· | ····················· | ···················· | ···················· | ···················· |
| L | 3514 | hCoV-19/England/ALDP-9EA81A/2020\|EPI_ISL_580033\|20 | ···················· | ···················· | ····················· | ···················· | ···················· | ···················· |
| L | 3515 | hCoV-19/England/ALDP-9E9DB2/2020\|EPI_ISL_580034\|20 | ···················· | ···················· | ····················· | ···················· | ···················· | ···················· |
| L | 3516 | hCoV-19/England/ALDP-9E9D67/2020\|EPI_ISL_580035\|20 | ···················· | ···················· | ····················· | ···················· | ···················· | ···················· |
| L | 3517 | hCoV-19/England/ALDP-9EAC78/2020\|EPI_ISL_580036\|20 | ···················· | ···················· | ····················· | ···················· | ···················· | ···················· |
| L | 3518 | hCoV-19/England/ALDP-9E7622/2020\|EPI_ISL_580037\|20 | ···················· | ···················· | ····················· | ···················· | ···················· | ···················· |
| L | 3519 | hCoV-19/England/ALDP-9EA285/2020\|EPI_ISL_580038\|20 | ···················· | ···················· | ····················· | ···················· | ···················· | ···················· |
| L | 3520 | hCoV-19/England/ALDP-9EABB7/2020\|EPI_ISL_580039\|20 | ···················· | ···················· | ····················· | ···················· | ···················· | ···················· |
| L | 3521 | hCoV-19/England/ALDP-9EB011/2020\|EPI_ISL_580040\|20 | ···················· | ···················· | ····················· | ···················· | ···················· | ···················· |
| L | 3522 | hCoV-19/England/ALDP-9EA4E9/2020\|EPI_ISL_580042\|20 | ···················· | ···················· | ····················· | ···················· | ···················· | ···················· |
| L | 3523 | hCoV-19/England/MILK-9E29B5/2020\|EPI_ISL_581300\|20 | ···················· | ···················· | ····················· | ···················· | ···················· | ··A················· |
| L | 3524 | hCoV-19/England/MILK-9E2F77/2020\|EPI_ISL_581301\|20 | ···················· | ···················· | ····················· | ···················· | ···················· | ···················· |
| L | 3525 | hCoV-19/England/MILK-9E2E98/2020\|EPI_ISL_581302\|20 | ···················· | ···················· | ····················· | ···················· | ···················· | ···················· |
| L | 3526 | hCoV-19/England/MILK-9E30A0/2020\|EPI_ISL_581303\|20 | ···················· | ···················· | ····················· | ···················· | ···················· | ···················· |
| L | 3527 | hCoV-19/England/MILK-9E2CE9/2020\|EPI_ISL_581305\|20 | ···················· | ···················· | ····················· | ···················· | ···················· | ···················· |
| L | 3528 | hCoV-19/Wales/MILK-9E2AA3/2020\|EPI_ISL_581306\|2020 | ···················· | ···················· | ····················· | ···················· | ···················· | ···················· |
| L | 3529 | hCoV-19/England/MILK-9E2A0D/2020\|EPI_ISL_581308\|20 | ···················· | ···················· | ····················· | ···················· | ···················· | ···················· |
| L | 3530 | hCoV-19/England/MILK-9E1FE1/2020\|EPI_ISL_581330\|20 | ···················· | ···················· | ····················· | ···················· | ···················· | ···················· |
| L | 3531 | hCoV-19/England/MILK-9E3134/2020\|EPI_ISL_581331\|20 | ···················· | ···················· | ····················· | ···················· | ···················· | ···················· |
| L | 3532 | hCoV-19/England/MILK-9E33D4/2020\|EPI_ISL_581332\|20 | ···················· | ···················· | ····················· | ···················· | ···················· | ···················· |
| L | 3533 | hCoV-19/England/MILK-9E32C8/2020\|EPI_ISL_581333\|20 | ···················· | ···················· | ····················· | ···················· | ···················· | ···················· |
| L | 3534 | hCoV-19/England/QEUH-9DE7EF/2020\|EPI_ISL_581334\|20 | ···················· | ···················· | ····················· | ···················· | ···················· | ···················· |
| L | 3535 | hCoV-19/Wales/MILK-9E1E20/2020\|EPI_ISL_581335\|2020 | ···················· | ···················· | ····················· | ···················· | ···················· | ···················· |
| L | 3536 | hCoV-19/England/MILK-9E20B0/2020\|EPI_ISL_581336\|20 | ···················· | ···················· | ····················· | ···················· | ···················· | ···················· |
| L | 3537 | hCoV-19/England/QEUH-9DE7FE/2020\|EPI_ISL_581337\|20 | ···················· | ···················· | ····················· | ···················· | ···················· | ···················· |
| L | 3538 | hCoV-19/England/MILK-9E31DA/2020\|EPI_ISL_581338\|20 | ···················· | ···················· | ····················· | ···················· | ···················· | ···················· |
| L | 3539 | hCoV-19/Wales/MILK-9E341D/2020\|EPI_ISL_581339\|2020 | ···················· | ···················· | ····················· | ···················· | ···················· | ···················· |
| L | 3540 | hCoV-19/England/MILK-9E1CAE/2020\|EPI_ISL_581340\|20 | ···················· | ···················· | ····················· | ···················· | ···················· | ···················· |
| L | 3541 | hCoV-19/England/QEUH-9DE6C4/2020\|EPI_ISL_581341\|20 | ···················· | ···················· | ····················· | ···················· | ···················· | ···················· |
| L | 3542 | hCoV-19/England/MILK-9E1C62/2020\|EPI_ISL_581342\|20 | ···················· | ···················· | ····················· | ···················· | ···················· | ···················· |
| L | 3543 | hCoV-19/Wales/MILK-9E2C43/2020\|EPI_ISL_581343\|2020 | ···················· | ···················· | ····················· | ···················· | ···················· | ···················· |
| L | 3544 | hCoV-19/England/QEUH-9DEC77/2020\|EPI_ISL_581344\|20 | ···················· | ···················· | ····················· | ···················· | ···················· | ···················· |
| L | 3545 | hCoV-19/England/MILK-9E2EF2/2020\|EPI_ISL_581345\|20 | ···················· | ···················· | ····················· | ···················· | ···················· | ···················· |
| L | 3546 | hCoV-19/England/QEUH-9DE59A/2020\|EPI_ISL_581346\|20 | ···················· | ·A·················· | ····················· | ···················· | ···················· | ··A················· |
| L | 3547 | hCoV-19/England/MILK-9E2FA4/2020\|EPI_ISL_581347\|20 | ···················· | ···················· | ····················· | ···················· | ·····T·············· | ···················· |
| L | 3548 | hCoV-19/England/MILK-9E2BCE/2020\|EPI_ISL_581348\|20 | ···················· | ···················· | ····················· | ···················· | ···················· | ···················· |
| L | 3549 | hCoV-19/England/MILK-9E2BEC/2020\|EPI_ISL_581349\|20 | ···················· | ···················· | ····················· | ···················· | ···················· | ···················· |
| L | 3550 | hCoV-19/England/MILK-9E2C70/2020\|EPI_ISL_581352\|20 | ···················· | ···················· | ····················· | ···················· | ···················· | ···················· |
| L | 3551 | hCoV-19/England/MILK-9E1EB7/2020\|EPI_ISL_581353\|20 | ···················· | ···················· | ····················· | ···················· | ···················· | ···················· |
| L | 3552 | hCoV-19/England/QEUH-9DEECC/2020\|EPI_ISL_581354\|20 | ···················· | ···················· | ····················· | ···················· | ···················· | ···················· |
| L | 3553 | hCoV-19/England/MILK-9E2627/2020\|EPI_ISL_581355\|20 | ···················· | ···················· | ····················· | ···················· | ···················· | ···················· |
| L | 3554 | hCoV-19/England/MILK-9E1F78/2020\|EPI_ISL_581356\|20 | ···················· | ···················· | ····················· | ···················· | ···················· | ···················· |
| L | 3555 | hCoV-19/England/MILK-9E1CBD/2020\|EPI_ISL_581357\|20 | ···················· | ···················· | ····················· | ···················· | ···················· | ···················· |
| L | 3556 | hCoV-19/England/QEUH-9DEEF9/2020\|EPI_ISL_581358\|20 | ···················· | ···················· | ····················· | ···················· | ···················· | ···················· |
| L | 3557 | hCoV-19/England/MILK-9E2D5F/2020\|EPI_ISL_581360\|20 | ···················· | ·A·················· | ····················· | ···················· | ···················· | ··A················· |
| L | 3558 | hCoV-19/Wales/MILK-9E328C/2020\|EPI_ISL_581361\|2020 | ···················· | ···················· | ····················· | ···················· | ···················· | ···················· |
| L | 3559 | hCoV-19/England/MILK-9E2BA0/2020\|EPI_ISL_581362\|20 | ···················· | ···················· | ····················· | ···················· | ···················· | ···················· |
| L | 3560 | hCoV-19/England/MILK-9E3170/2020\|EPI_ISL_581363\|20 | ···················· | ···················· | ····················· | ···················· | ···················· | ···················· |
| L | 3561 | hCoV-19/England/MILK-9E1CDB/2020\|EPI_ISL_581364\|20 | ···················· | ·A·················· | ····················· | ···················· | ···················· | ··A················· |
| L | 3562 | hCoV-19/England/MILK-9E33B6/2020\|EPI_ISL_581365\|20 | ···················· | C··················· | ····················· | ···················· | ···················· | ···················· |
| L | 3563 | hCoV-19/England/ALDP-9CB23F/2020\|EPI_ISL_581367\|20 | ···················· | ···················· | ····················· | ···················· | ···················· | ···················· |
| L | 3564 | hCoV-19/England/MILK-976400/2020\|EPI_ISL_581368\|20 | ···················· | ···················· | ····················· | ···················· | ···················· | ···················· |
| L | 3565 | hCoV-19/England/MILK-974930/2020\|EPI_ISL_581369\|20 | ···················· | ···················· | ····················· | ···················· | ···················· | ···················· |

```
L    3566 hCoV-19/England/MILK-974A4C/2020|EPI_ISL_581370|20    ....................  ....................  ....................  ....................  .....T..............  ....................
L    3567 hCoV-19/England/MILK-9D3F2E/2020|EPI_ISL_581371|20    ....................  ....................  ....................  ....................  ....................  ....................
L    3568 hCoV-19/England/MILK-9D4725/2020|EPI_ISL_581372|20    ....................  ....................  ....................  ....................  ....................  ....................
L    3569 hCoV-19/England/MILK-9D4734/2020|EPI_ISL_581373|20    ....................  ....................  ....................  ....................  ....................  ....................
L    3570 hCoV-19/United Arab Emirates/skmc-920568379/2020|E    ....................  ....................  ....................  ....................  ....................  ....................
L    3571 hCoV-19/United Arab Emirates/skmc-004/2020|EPI_ISL    ....................  ....................  ....................  ....................  ....................  ....................
L    3572 hCoV-19/United Arab Emirates/skmc-227072/2020|EPI_    ....................  ....................  ....................  ....................  ....................  ....................
L    3573 hCoV-19/United Arab Emirates/skmc-3580730/2020|EPI    ....................  ....................  ....................  ....................  ....................  ....................
L    3574 hCoV-19/United Arab Emirates/skmc-3759960/2020|EPI    ....................  ....................  ....................  ....................  ....................  ....................
L    3575 hCoV-19/United Arab Emirates/skmc-3819699/2020|EPI    ....................  ....................  ....................  ....................  ....................  ....................
L    3576 hCoV-19/United Arab Emirates/skmc-3819883/2020|EPI    ....................  ....................  ....................  ....................  ....................  ....................
L    3577 hCoV-19/United Arab Emirates/skmc-3883762/2020|EPI    ....................  ....................  ....................  ....................  ....................  ....................
L    3578 hCoV-19/United Arab Emirates/skmc-3890804/2020|EPI    ....................  ....................  ....................  ....................  ....................  ....................
L    3579 hCoV-19/United Arab Emirates/skmc-3890161/2020|EPI    ....................  ....................  ....................  ....................  ....................  ....................
L    3580 hCoV-19/United Arab Emirates/skmc-3891274/2020|EPI    ....................  ....................  ....................  ....................  ....................  ....................
L    3581 hCoV-19/United Arab Emirates/skmc-3942334/2020|EPI    ....................  ....................  ....................  ....................  ....................  ....................
L    3582 hCoV-19/Italy/SIC-IZSBM169-56/2020|EPI_ISL_582692|    ....................  ....................  ....................  ....................  ....................  ....................
L    3583 hCoV-19/Italy/SIC-IZSBM169-70/2020|EPI_ISL_582693|    ....................  ....................  ....................  ....................  ....................  ....................
L    3584 hCoV-19/Italy/SIC-IZSBM169-76/2020|EPI_ISL_582694|    ....................  ....................  ....................  ....................  ....................  ....................
L    3585 hCoV-19/Wales/PHWC-16D0F1/2020|EPI_ISL_574202|2020    ....................  ....................  ....................  ....................  ....................  ....................
L    3586 hCoV-19/Wales/PHWC-16D10D/2020|EPI_ISL_574203|2020    ....................  ....................  ....................  ....................  ....................  ....................
L    3587 hCoV-19/Wales/PHWC-16D11C/2020|EPI_ISL_574204|2020    ....................  ....................  ....................  ....................  ....................  ....................
L    3588 hCoV-19/Wales/PHWC-16D12B/2020|EPI_ISL_574205|2020    ....................  ....................  ....................  ....................  ....................  ....................
L    3590 hCoV-19/Wales/PHWC-16D176/2020|EPI_ISL_574207|2020    ....................  ....................  ....................  ....................  ....................  ....................
L    3591 hCoV-19/Wales/PHWC-16D1B2/2020|EPI_ISL_574208|2020    ....................  ....................  ....................  ....................  ....................  ....................
L    3593 hCoV-19/Wales/PHWC-16D1D0/2020|EPI_ISL_574210|2020    ....................  ....................  ....................  ....................  ....................  ....................
L    3598 hCoV-19/Wales/PHWC-16D237/2020|EPI_ISL_574215|2020    ....................  ....................  ....................  ....................  ....................  ....................
L    3599 hCoV-19/Wales/PHWC-16D246/2020|EPI_ISL_574216|2020    ....................  ....................  ....................  ....................  ....................  ....................
L    3600 hCoV-19/Wales/PHWC-16D255/2020|EPI_ISL_574217|2020    ....................  ....................  ....................  ....................  ....................  ....................
L    3602 hCoV-19/Wales/PHWC-16D273/2020|EPI_ISL_574219|2020    ....................  ....................  ....................  ....................  ....................  ....................
L    3603 hCoV-19/Wales/PHWC-16D282/2020|EPI_ISL_574220|2020    ....................  ....................  ....................  ....................  ....................  ....................
L    3604 hCoV-19/Wales/PHWC-16D2A0/2020|EPI_ISL_574221|2020    ....................  ....................  ....................  ....................  ....................  ....................
L    3605 hCoV-19/Wales/PHWC-16D2DD/2020|EPI_ISL_574223|2020    ....................  ....................  ....................  ....................  ....................  ....................
L    3606 hCoV-19/Wales/PHWC-16D2EC/2020|EPI_ISL_574224|2020    ....................  ....................  ....................  ....................  ....................  ....................
L    3607 hCoV-19/Wales/PHWC-16D2FB/2020|EPI_ISL_574225|2020    ....................  ....................  ....................  ....................  ....................  ....................
L    3609 hCoV-19/Wales/PHWC-16D361/2020|EPI_ISL_574227|2020    ....................  ....................  ....................  ....................  NNN.................  ....................    1
L    3611 hCoV-19/Wales/PHWC-16D38F/2020|EPI_ISL_574229|2020    ....................  ....................  ....................  ....................  ....................  ....................
L    3613 hCoV-19/Wales/PHWC-16D3AD/2020|EPI_ISL_574231|2020    ....................  ....................  ....................  ....................  ....................  ....................
L    3614 hCoV-19/Wales/PHWC-16D3DA/2020|EPI_ISL_574232|2020    ....................  ....................  ....................  ....................  ....................  ....................
L    3615 hCoV-19/Wales/PHWC-16D3E9/2020|EPI_ISL_574233|2020    ....................  ....................  ....................  ....................  ....................  ....................
L    3616 hCoV-19/Wales/PHWC-16D3F8/2020|EPI_ISL_574234|2020    ....................  ....................  ....................  ....................  ....................  ....................
L    3618 hCoV-19/Wales/PHWC-16D422/2020|EPI_ISL_574236|2020    ....................  ....................  ....................  ....................  ....................  ....................
L    3619 hCoV-19/Wales/PHWC-16D440/2020|EPI_ISL_574237|2020    ....................  ....................  ....................  ....................  ....................  ....................
L    3620 hCoV-19/Wales/PHWC-16D45F/2020|EPI_ISL_574238|2020    ....................  ....................  ....................  ....................  ....................  ....................
L    3621 hCoV-19/Wales/PHWC-16D46E/2020|EPI_ISL_574239|2020    ....................  ....................  ....................  ....................  ....................  ....................
L    3622 hCoV-19/Wales/PHWC-16D48C/2020|EPI_ISL_574240|2020    ....................  ....................  ....................  ....................  ....................  ....................
L    3624 hCoV-19/Wales/PHWC-16D4AA/2020|EPI_ISL_574242|2020    ....................  ....................  ....................  ....................  ....................  ....................
L    3625 hCoV-19/Wales/PHWC-16D4B9/2020|EPI_ISL_574243|2020    ....................  ....................  ....................  ....................  ....................  ....................
L    3627 hCoV-19/Wales/PHWC-16D4E6/2020|EPI_ISL_574245|2020    ....................  ....................  ....................  ....................  ....................  ....................
L    3628 hCoV-19/Wales/PHWC-16D4F5/2020|EPI_ISL_574246|2020    ....................  ....................  ....................  ....................  ....................  ....................
L    3629 hCoV-19/Wales/PHWC-16D501/2020|EPI_ISL_574247|2020    ....................  ....................  ....................  ....................  ....................  ....................
L    3632 hCoV-19/Wales/PHWC-16D55C/2020|EPI_ISL_574250|2020    ....................  ....................  ....................  ....................  ....................  ....................
L    3633 hCoV-19/Wales/PHWC-16D56B/2020|EPI_ISL_574251|2020    ....................  ....................  ....................  ....................  ....................  ....................
L    3634 hCoV-19/Wales/PHWC-16D57A/2020|EPI_ISL_574252|2020    ....................  ....................  ....................  ....................  ....................  ....................
L    3635 hCoV-19/Wales/PHWC-16D589/2020|EPI_ISL_574253|2020    ....................  ....................  ....................  ....................  ....................  ....................
L    3636 hCoV-19/Wales/PHWC-16D598/2020|EPI_ISL_574254|2020    ....................  ....................  ....................  ....................  ....................  ....................
L    3638 hCoV-19/Wales/PHWC-16D5C5/2020|EPI_ISL_574256|2020    ....................  ....................  ....................  ....................  ....................  ....................
L    3639 hCoV-19/Wales/QEUH-9BA2E8/2020|EPI_ISL_574257|2020    ....................  ....................  ....................  ....................  ....................  ....................
L    3640 hCoV-19/Wales/QEUH-9D16EF/2020|EPI_ISL_574258|2020    ....................  ....................  ....................  ....................  ....................  ....................
L    3641 hCoV-19/Germany/NW-HHU-D872/2020|EPI_ISL_574259|20    ....................  ....................  ....................  ....................  ....................  ....................
L    3642 hCoV-19/USA/NM-DOH-2020312737/2020|EPI_ISL_574260|    ....................  ....................  ....................  ....................  ....................  ....................
L    3643 hCoV-19/USA/NM-DOH-2020313047/2020|EPI_ISL_574261|    ....................  ....................  ....................  ....................  ....................  ....................
L    3644 hCoV-19/USA/NM-DOH-2020313085/2020|EPI_ISL_574262|    ....................  ....................  ....................  ....................  ....................  ....................
L    3645 hCoV-19/USA/NM-DOH-2020313106/2020|EPI_ISL_574263|    ....................  ....................  ....................  ....................  ....................  ....................
L    3646 hCoV-19/USA/NM-DOH-2020313186/2020|EPI_ISL_574264|    ....................  .A..................  ....................  ....................  ....................  ....................
L    3647 hCoV-19/USA/NM-DOH-2020313196/2020|EPI_ISL_574265|    ....................  ....................  ....................  ....................  ....................  ....................
L    3648 hCoV-19/USA/NM-DOH-2020313239/2020|EPI_ISL_574266|    ....................  ....................  ....................  ....................  ....................  ....................
L    3649 hCoV-19/USA/NM-DOH-2020313314/2020|EPI_ISL_574267|    ....................  ....A...............  ....................  ....................  ....................  ....................
L    3650 hCoV-19/USA/NM-DOH-2020313350/2020|EPI_ISL_574268|    ....................  ....................  ....................  ....................  ....................  ....................
L    3651 hCoV-19/USA/NM-DOH-2020313398/2020|EPI_ISL_574269|    ....................  .A..................  ....................  ....................  ....................  ....................
L    3652 hCoV-19/USA/NM-DOH-2020313467/2020|EPI_ISL_574270|    ....................  ....................  ....................  ....................  ....................  ....................
L    3653 hCoV-19/USA/NM-DOH-2020313553/2020|EPI_ISL_574271|    ....................  ....................  ....................  ....................  ....................  ....................
L    3654 hCoV-19/USA/NM-DOH-2020313623/2020|EPI_ISL_574272|    ....................  ....A...............  ....................  ....................  ....................  ....................
L    3655 hCoV-19/USA/NM-DOH-2020313625/2020|EPI_ISL_574273|    ....................  ....................  ....................  ....................  ....................  ....................
L    3656 hCoV-19/USA/NM-DOH-2020313776/2020|EPI_ISL_574274|    ....................  ....................  ....................  ....................  ....................  ....................
L    3657 hCoV-19/USA/NM-DOH-2020314766/2020|EPI_ISL_574275|    ....................  ....................  ....................  ....................  ....................  ....................
L    3658 hCoV-19/USA/NM-DOH-2020314787/2020|EPI_ISL_574276|    ....................  ..............A.....  ....................  ....................  ....................  ....................
L    3659 hCoV-19/USA/NM-DOH-2020314819/2020|EPI_ISL_574277|    ....................  ....................  ....................  ....................  ....................  ....................
L    3660 hCoV-19/USA/NM-DOH-2020314838/2020|EPI_ISL_574278|    ....................  ....................  ....................  ....................  ....................  ....................
L    3661 hCoV-19/USA/NM-DOH-2020314841/2020|EPI_ISL_574279|    ....................  ....................  ....................  ....................  ....................  ....................
```

L 3662 hCoV-19/USA/NM-DOH-2020314850/2020|EPI_ISL_574280| ···················· ···················· ····················· ···················· ···················· ····················
L 3663 hCoV-19/USA/NM-DOH-2020314875/2020|EPI_ISL_574281| ···················· ···················· ····················· ···················· ···················· ····················
L 3664 hCoV-19/USA/NM-DOH-2020314888/2020|EPI_ISL_574282| ···················· ···················· ····················· ···················· ···················· ····················
L 3665 hCoV-19/USA/NM-DOH-2020314889/2020|EPI_ISL_574283| ···················· ···················· ····················· ···················· ···················· ····················
L 3666 hCoV-19/USA/NM-DOH-2020315014/2020|EPI_ISL_574284| ···················· ···················· ····················· ···················· ···················· ····················
L 3667 hCoV-19/USA/NM-DOH-2020315026/2020|EPI_ISL_574285| ···················· ···A················ ····················· ···················· ···················· ····················
L 3668 hCoV-19/USA/NM-DOH-2020315299/2020|EPI_ISL_574286| ···················· ···················· ····················· ···················· ···················· ····················
L 3669 hCoV-19/USA/NM-DOH-2020315301/2020|EPI_ISL_574287| ···················· ···················· ····················· ···················· ···················· ····················
L 3670 hCoV-19/England/ALDP-9CAA29/2020|EPI_ISL_575533|20 ···················· ···················· ····················· ···················· ···················· ····················
L 3671 hCoV-19/England/ALDP-9CB6F7/2020|EPI_ISL_575534|20 ···················· ···················· ····················· ···················· ···················· ····················
L 3672 hCoV-19/England/ALDP-9CAF2A/2020|EPI_ISL_575535|20 ···················· ···················· ····················· ···················· ···················· ····················
L 3673 hCoV-19/England/ALDP-9CBCD7/2020|EPI_ISL_575537|20 ···················· ···················· ····················· ···················· ···················· ····················
L 3674 hCoV-19/England/ALDP-9CAA83/2020|EPI_ISL_575538|20 ···················· ···················· ····················· ···················· ···················· ····················
L 3675 hCoV-19/England/ALDP-9CAC23/2020|EPI_ISL_575539|20 ···················· ·A·················· ····················· ···················· ···················· ··A·················
L 3676 hCoV-19/England/ALDP-9CBA82/2020|EPI_ISL_575540|20 ···················· ·A·················· ····················· ···················· ···················· ····················
L 3677 hCoV-19/England/ALDP-9CB484/2020|EPI_ISL_575541|20 ···················· ···················· ····················· ···················· ···················· ····················
L 3678 hCoV-19/England/ALDP-9CAB08/2020|EPI_ISL_575542|20 ···················· ···················· ····················· ···················· ···················· ····················
L 3679 hCoV-19/England/ALDP-9CBC22/2020|EPI_ISL_575544|20 ···················· ···················· ····················· ···················· ···················· ····················
L 3680 hCoV-19/England/ALDP-9CB7F4/2020|EPI_ISL_575545|20 ···················· ···················· ····················· ···················· ···················· ····················
L 3681 hCoV-19/England/ALDP-9CAC7E/2020|EPI_ISL_575546|20 ···················· ···················· ····················· ···················· ···················· ····················
L 3682 hCoV-19/England/ALDP-9CC025/2020|EPI_ISL_575547|20 ···················· ···················· ····················· ···················· ···················· ····················
L 3683 hCoV-19/England/ALDP-9CC070/2020|EPI_ISL_575548|20 ···················· ···················· ····················· ···················· ···················· ····················
L 3684 hCoV-19/England/ALDP-9CB651/2020|EPI_ISL_575549|20 ···················· ···················· ····················· ···················· ···················· ····················
L 3685 hCoV-19/England/ALDP-9CB1C9/2020|EPI_ISL_575551|20 ···················· ···················· ····················· ···················· ···················· ····················
L 3686 hCoV-19/England/ALDP-9CAF66/2020|EPI_ISL_575555|20 ···················· ·A·················· ····················· ···················· ···················· ····················
L 3687 hCoV-19/England/ALDP-9CBFA1/2020|EPI_ISL_575556|20 ···················· ···················· ····················· ···················· ···················· ····················
L 3688 hCoV-19/England/ALDP-9CB053/2020|EPI_ISL_575557|20 ···················· ···················· ····················· ···················· ···················· ····················
L 3689 hCoV-19/England/ALDP-9CB590/2020|EPI_ISL_575558|20 ···················· ···················· ····················· ···················· ···················· ····················
L 3690 hCoV-19/England/ALDP-9CBA55/2020|EPI_ISL_575559|20 ···················· ···················· ····················· ···················· ···················· ····················
L 3691 hCoV-19/England/ALDP-9CB985/2020|EPI_ISL_575560|20 ···················· ···················· ····················· ···················· ···················· ····················
L 3692 hCoV-19/England/ALDP-9CB09F/2020|EPI_ISL_575562|20 ·········T·········· ···················· ····················· ···················· ···················· ····················
L 3693 hCoV-19/England/ALDP-9CBFDE/2020|EPI_ISL_575563|20 ···················· ···················· ····················· ···················· ···················· ····················
L 3694 hCoV-19/England/ALDP-9CB19C/2020|EPI_ISL_575564|20 ···················· ···················· ····················· ···················· ···················· ····················
L 3695 hCoV-19/England/ALDP-9CB8D3/2020|EPI_ISL_575565|20 ···················· ···················· ····················· ···················· ···················· ····················
L 3696 hCoV-19/England/ALDP-9CAAB0/2020|EPI_ISL_575566|20 ···················· ···················· ····················· ···················· ···················· ····················
L 3697 hCoV-19/England/ALDP-9CAA47/2020|EPI_ISL_575567|20 ···················· ···················· ····················· ···················· ···················· ····················
L 3698 hCoV-19/England/ALDP-9CB563/2020|EPI_ISL_575568|20 ···················· ···················· ····················· ···················· ···················· ····················
L 3699 hCoV-19/England/ALDP-9CBF56/2020|EPI_ISL_575569|20 ···················· ···················· ····················· ···················· ···················· ····················
L 3700 hCoV-19/England/ALDP-9CB2A8/2020|EPI_ISL_575570|20 ···················· ·A·················· ····················· ···················· ···················· ··A·················
L 3701 hCoV-19/England/ALDP-9CB299/2020|EPI_ISL_575571|20 ···················· ·A·················· ····················· ···················· ···················· ··A·················
L 3702 hCoV-19/England/ALDP-9CAEC3/2020|EPI_ISL_575572|20 ···················· ···················· ····················· ···················· ···················· ····················
L 3703 hCoV-19/England/ALDP-9CBB70/2020|EPI_ISL_575573|20 ···················· ···················· ····················· ···················· ···················· ····················
L 3704 hCoV-19/England/ALDP-9CB615/2020|EPI_ISL_575574|20 ···················· ···················· ····················· ···················· ···················· ····················
L 3705 hCoV-19/England/ALDP-9CBDE3/2020|EPI_ISL_575575|20 ···················· ···················· ····················· ···················· ···················· ····················
L 3706 hCoV-19/England/ALDP-9CAE1E/2020|EPI_ISL_575576|20 ···················· ···················· ····················· ···················· ···················· ····················
L 3707 hCoV-19/England/ALDP-9CB017/2020|EPI_ISL_575577|20 ···················· ···················· ····················· ···················· ···················· ····················
L 3708 hCoV-19/England/ALDP-9CB0BD/2020|EPI_ISL_575578|20 ···················· ···················· ····················· ···················· ···················· ····················
L 3709 hCoV-19/England/ALDP-9CB554/2020|EPI_ISL_575580|20 ···················· ···················· ····················· ···················· ···················· ····················
L 3710 hCoV-19/England/ALDP-9CADD5/2020|EPI_ISL_575582|20 ···················· ···················· ····················· ···················· ···················· ····················
L 3711 hCoV-19/England/ALDP-9CAA65/2020|EPI_ISL_575584|20 ···················· ···················· ····················· ···················· ···················· ····················
L 3712 hCoV-19/England/ALDP-9CB545/2020|EPI_ISL_575585|20 ···················· ···················· ····················· ···················· ···················· ····················
L 3713 hCoV-19/England/ALDP-9CB132/2020|EPI_ISL_575586|20 ···················· ···················· ····················· ···················· ···················· ····················
L 3714 hCoV-19/England/ALDP-9CAC41/2020|EPI_ISL_575587|20 ···················· ···················· ····················· ···················· ···················· ····················
L 3715 hCoV-19/England/ALDP-9CB3D2/2020|EPI_ISL_575588|20 ···················· ···················· ····················· ···················· ···················· ····················
L 3716 hCoV-19/England/ALDP-9CB3C3/2020|EPI_ISL_575589|20 ···················· ···················· ····················· ···················· ···················· ····················
L 3717 hCoV-19/England/ALDP-9CAFDF/2020|EPI_ISL_575590|20 ···················· ·A·················· ····················· ···················· ···················· ····················
L 3718 hCoV-19/England/ALDP-9CBD89/2020|EPI_ISL_575591|20 ···················· ···················· ····················· ···················· ···················· ····················
L 3719 hCoV-19/England/ALDP-9CB9B2/2020|EPI_ISL_575592|20 ···················· ···················· ····················· ···················· ···················· ····················
L 3720 hCoV-19/England/ALDP-9CAF57/2020|EPI_ISL_575593|20 ···················· ···················· ····················· ···················· ···················· ····················
L 3721 hCoV-19/England/ALDP-9CBD6B/2020|EPI_ISL_575594|20 ···················· ···················· ····················· ···················· ···················· ····················
L 3722 hCoV-19/England/ALDP-9CBA28/2020|EPI_ISL_575595|20 ···················· ···················· ····················· ···················· ···················· ····················
L 3723 hCoV-19/England/ALDP-9CB5EB/2020|EPI_ISL_575596|20 ···················· ···················· ····················· ···················· ···················· ····················
L 3724 hCoV-19/England/ALDP-9CBCF5/2020|EPI_ISL_575597|20 ···················· ···················· ····················· ···················· ···················· ····················
L 3725 hCoV-19/England/ALDP-9CB79A/2020|EPI_ISL_575599|20 ···················· ·A·················· ····················· ···················· ···················· ··A·················
L 3726 hCoV-19/England/ALDP-9D9BE8/2020|EPI_ISL_576801|20 ···················· ···················· ····················· ···················· ···················· ····················
L 3727 hCoV-19/England/ALDP-9D9C5E/2020|EPI_ISL_576806|20 ···················· ···················· ····················· ···················· ···················· ····················
L 3728 hCoV-19/England/ALDP-9D9C6D/2020|EPI_ISL_576807|20 ···················· ···················· ····················· ···················· ···················· ····················
L 3729 hCoV-19/England/ALDP-9D9C7C/2020|EPI_ISL_576808|20 ···················· ···················· ····················· ···················· ···················· ····················
L 3730 hCoV-19/England/ALDP-9D9CB8/2020|EPI_ISL_576810|20 ···················· ···················· ····················· ···················· ···················· ····················
L 3731 hCoV-19/England/ALDP-9D9D00/2020|EPI_ISL_576811|20 ···················· ···················· ····················· ···················· ······T············· ····················
L 3732 hCoV-19/England/ALDP-9D9D1F/2020|EPI_ISL_576812|20 ···················· ···················· ····················· ···················· ···················· ····················
L 3733 hCoV-19/England/ALDP-9D9D2E/2020|EPI_ISL_576813|20 ···················· ···················· ····················· ···················· ···················· ····················
L 3734 hCoV-19/England/ALDP-9D9D88/2020|EPI_ISL_576814|20 ···················· ···················· ····················· ···················· ···················· ····················
L 3735 hCoV-19/England/ALDP-9D9DA6/2020|EPI_ISL_576815|20 ···················· ···················· ····················· ···················· ···················· ····················
L 3736 hCoV-19/England/ALDP-9D9DC4/2020|EPI_ISL_576816|20 ···················· ···················· ····················· ···················· ···················· ····················
L 3737 hCoV-19/England/ALDP-9D9DE2/2020|EPI_ISL_576818|20 ···················· ···················· ····················· ···················· ···················· ····················
L 3738 hCoV-19/England/ALDP-9D9DF1/2020|EPI_ISL_576819|20 ···················· ···················· ····················· ···················· ···················· ····················
L 3739 hCoV-19/England/ALDP-9DA349/2020|EPI_ISL_576864|20 ···················· ···················· ····················· ···················· ···················· ····················
L 3740 hCoV-19/England/ALDP-9DA358/2020|EPI_ISL_576865|20 ···················· ···················· ····················· ···················· ···················· ····················
L 3741 hCoV-19/England/ALDP-9DA385/2020|EPI_ISL_576867|20 ···················· ···················· ····················· ···················· ···················· ····················

| | | | | | | | |
|---|---|---|---|---|---|---|---|
| L | 3742 hCoV-19/England/ALDP-9DA3A3/2020\|EPI_ISL_576868\|20 | ···················· | ···················· | ····················· | ···················· | ···················· | ···················· |
| L | 3743 hCoV-19/England/ALDP-9DA3B2/2020\|EPI_ISL_576869\|20 | ···················· | ···················· | ····················· | ···················· | ···················· | ···················· |
| L | 3744 hCoV-19/England/ALDP-9DA3C1/2020\|EPI_ISL_576870\|20 | ···················· | ·A·················· | ····················· | ···················· | ···················· | ···A················ |
| L | 3745 hCoV-19/England/ALDP-9DA419/2020\|EPI_ISL_576871\|20 | ···················· | ···················· | ····················· | ···················· | ···················· | ···················· |
| L | 3746 hCoV-19/England/ALDP-9DA437/2020\|EPI_ISL_576872\|20 | ···················· | ···················· | ····················· | ···················· | ···················· | ···················· |
| L | 3747 hCoV-19/England/ALDP-9DA455/2020\|EPI_ISL_576874\|20 | ···················· | ···················· | ····················· | ···················· | ···················· | ···················· |
| L | 3748 hCoV-19/England/ALDP-9DA464/2020\|EPI_ISL_576875\|20 | ···················· | ···················· | ····················· | ···················· | ···················· | ···················· |
| L | 3749 hCoV-19/England/ALDP-9DA4A0/2020\|EPI_ISL_576876\|20 | ···················· | ···················· | ····················· | ···················· | ···················· | ···················· |
| L | 3752 hCoV-19/England/BIRM-63D9B/2020\|EPI_ISL_576883\|202 | ···················· | ···················· | ····················· | ···················· | ···················· | ···················· |
| L | 3753 hCoV-19/England/BIRM-63DAA/2020\|EPI_ISL_576884\|202 | ···················· | ···················· | ····················· | ···················· | ···················· | ···················· |
| L | 3754 hCoV-19/England/BIRM-63E10/2020\|EPI_ISL_576887\|202 | ···················· | ···················· | ····················· | ···················· | ···················· | ···················· |
| L | 3755 hCoV-19/England/BIRM-63E2F/2020\|EPI_ISL_576888\|202 | ···················· | ···················· | ····················· | ···················· | ···················· | ···················· |
| L | 3756 hCoV-19/England/BIRM-63E3E/2020\|EPI_ISL_576889\|202 | ···················· | ···················· | ····················· | ···················· | ···················· | ···················· |
| L | 3757 hCoV-19/England/BIRM-63E4D/2020\|EPI_ISL_576890\|202 | ···················· | ···················· | ····················· | ···················· | ···················· | ···················· |
| L | 3758 hCoV-19/England/BIRM-63E89/2020\|EPI_ISL_576892\|202 | ···················· | ···················· | ····················· | ···················· | ···················· | ···················· |
| L | 3759 hCoV-19/England/BIRM-63E98/2020\|EPI_ISL_576893\|202 | ···················· | ···················· | ····················· | ···················· | ···················· | ···················· |
| L | 3760 hCoV-19/England/BIRM-63EA7/2020\|EPI_ISL_576894\|202 | ···················· | ···················· | ····················· | ···················· | ···················· | ···················· |
| L | 3762 hCoV-19/New Zealand/20CV0265/2020\|EPI_ISL_579060\|2 | ···················· | ···················· | ····················· | ···················· | ···················· | ···················· |
| L | 3763 hCoV-19/New Zealand/20CV0266/2020\|EPI_ISL_579061\|2 | ···················· | ···················· | ····················· | ···················· | ···················· | ···················· |
| L | 3764 hCoV-19/New Zealand/20CV0269/2020\|EPI_ISL_579063\|2 | ···················· | ···················· | ····················· | ···················· | ···················· | ···················· |
| L | 3765 hCoV-19/New Zealand/20CV0270/2020\|EPI_ISL_579064\|2 | ···················· | ···················· | ····················· | ···················· | ···················· | ···················· |
| L | 3766 hCoV-19/New Zealand/20CV0271/2020\|EPI_ISL_579065\|2 | ···················· | ···················· | ····················· | ···················· | ···················· | ···················· |
| L | 3767 hCoV-19/New Zealand/20CV0273/2020\|EPI_ISL_579066\|2 | ···················· | ···················· | ····················· | ···················· | ···················· | ···················· |
| L | 3768 hCoV-19/New Zealand/20CV0274/2020\|EPI_ISL_579067\|2 | ···················· | ···················· | ····················· | ···················· | ···················· | ···················· |
| L | 3769 hCoV-19/New Zealand/20CV0276/2020\|EPI_ISL_579068\|2 | ···················· | ···················· | ····················· | ···················· | ···················· | ···················· |
| L | 3770 hCoV-19/New Zealand/20CV0277/2020\|EPI_ISL_579069\|2 | ···················· | ···················· | ····················· | ···················· | ···················· | ···················· |
| L | 3771 hCoV-19/New Zealand/20CV0278/2020\|EPI_ISL_579070\|2 | ···················· | ···················· | ····················· | ···················· | ···················· | ···················· |
| L | 3772 hCoV-19/New Zealand/20CV0280/2020\|EPI_ISL_579071\|2 | ···················· | ···················· | ····················· | ···················· | ···················· | ···················· |
| L | 3773 hCoV-19/New Zealand/20CV0281/2020\|EPI_ISL_579072\|2 | ···················· | ···················· | ····················· | ···················· | ···················· | ···················· |
| L | 3774 hCoV-19/New Zealand/20CV0283/2020\|EPI_ISL_579073\|2 | ···················· | ···················· | ····················· | ···················· | ···················· | ···················· |
| L | 3775 hCoV-19/New Zealand/20CV0284/2020\|EPI_ISL_579074\|2 | ···················· | ···················· | ····················· | ···················· | ···················· | ···················· |
| L | 3776 hCoV-19/New Zealand/20CV0285/2020\|EPI_ISL_579075\|2 | ···················· | ···················· | ····················· | ···················· | ···················· | ···················· |
| L | 3777 hCoV-19/New Zealand/20CV0287/2020\|EPI_ISL_579077\|2 | ···················· | ···················· | ····················· | ···················· | ···················· | ···················· |
| L | 3778 hCoV-19/New Zealand/20CV0288/2020\|EPI_ISL_579078\|2 | ···················· | ···················· | ····················· | ···················· | ···················· | ···················· |
| L | 3779 hCoV-19/New Zealand/20CV0289/2020\|EPI_ISL_579079\|2 | ···················· | ···················· | ····················· | ···················· | ···················· | ···················· |
| L | 3780 hCoV-19/New Zealand/20CV0290/2020\|EPI_ISL_579080\|2 | ···················· | ···················· | ····················· | ···················· | ···················· | ···················· |
| L | 3781 hCoV-19/New Zealand/20CV0291/2020\|EPI_ISL_579081\|2 | ···················· | ···················· | ····················· | ···················· | ···················· | ···················· |
| L | 3782 hCoV-19/New Zealand/20CV0292/2020\|EPI_ISL_579082\|2 | ···················· | ···················· | ····················· | ···················· | ···················· | ···················· |
| L | 3783 hCoV-19/New Zealand/20CV0293/2020\|EPI_ISL_579083\|2 | ···················· | ···················· | ····················· | ···················· | ···················· | ···················· |
| L | 3784 hCoV-19/New Zealand/20CV0294/2020\|EPI_ISL_579084\|2 | ···················· | ···················· | ····················· | ···················· | ···················· | ···················· |
| L | 3785 hCoV-19/New Zealand/20CV0295/2020\|EPI_ISL_579085\|2 | ···················· | ···················· | ····················· | ···················· | ···················· | ···················· |
| L | 3788 hCoV-19/New Zealand/20CV0350/2020\|EPI_ISL_579099\|2 | ···················· | ·A·················· | ····················· | ···················· | ···················· | ···A················ |
| L | 3789 hCoV-19/England/ALDP-9E9ECE/2020\|EPI_ISL_580043\|20 | ···················· | ···················· | ····················· | ···················· | ···················· | ···················· |
| L | 3790 hCoV-19/England/ALDP-9EA698/2020\|EPI_ISL_580044\|20 | ···················· | ···················· | ····················· | ···················· | ···················· | ···················· |
| L | 3791 hCoV-19/England/ALDP-9EAC96/2020\|EPI_ISL_580045\|20 | ···················· | ···················· | ····················· | ···················· | ···················· | ···················· |
| L | 3792 hCoV-19/England/ALDP-9EAB6C/2020\|EPI_ISL_580046\|20 | ···················· | ···················· | ····················· | ···················· | ···················· | ···················· |
| L | 3793 hCoV-19/England/ALDP-9E7640/2020\|EPI_ISL_580047\|20 | ···················· | ···················· | ····················· | ···················· | ···················· | ···················· |
| L | 3794 hCoV-19/England/ALDP-9E9CB5/2020\|EPI_ISL_580048\|20 | ···················· | ···················· | ····················· | ···················· | ···················· | ···················· |
| L | 3795 hCoV-19/England/ALDP-9E9FAD/2020\|EPI_ISL_580049\|20 | ···················· | ···················· | ····················· | ···················· | ···················· | ···················· |
| L | 3796 hCoV-19/England/ALDP-9E9FBC/2020\|EPI_ISL_580050\|20 | ···················· | ···················· | ····················· | ···················· | ···················· | ···················· |
| L | 3797 hCoV-19/England/ALDP-9E9CD3/2020\|EPI_ISL_580051\|20 | ···················· | ···················· | ····················· | ···················· | ···················· | ···················· |
| L | 3798 hCoV-19/England/ALDP-9EA540/2020\|EPI_ISL_580052\|20 | ···················· | ·A·················· | ····················· | ···················· | ···················· | ···················· |
| L | 3799 hCoV-19/England/ALDP-9EA382/2020\|EPI_ISL_580053\|20 | ···················· | ···················· | ····················· | ···················· | ···················· | ···················· |
| L | 3800 hCoV-19/England/ALDP-9EAECD/2020\|EPI_ISL_580055\|20 | ···················· | ···················· | ····················· | ···················· | ···················· | ···················· |
| L | 3801 hCoV-19/England/ALDP-9EA62F/2020\|EPI_ISL_580056\|20 | ···················· | ···················· | ····················· | ···················· | ···················· | ···················· |
| L | 3802 hCoV-19/England/ALDP-9EA434/2020\|EPI_ISL_580057\|20 | ···················· | ···················· | ····················· | ···················· | ···················· | ···················· |
| L | 3803 hCoV-19/England/ALDP-9EA56E/2020\|EPI_ISL_580058\|20 | ···················· | ···················· | ····················· | ···················· | ···················· | ···················· |
| L | 3804 hCoV-19/England/ALDP-9EA5D7/2020\|EPI_ISL_580059\|20 | ···················· | ···················· | ····················· | ···················· | ···················· | ···················· |
| L | 3805 hCoV-19/England/ALDP-9EA407/2020\|EPI_ISL_580063\|20 | ···················· | ···················· | ····················· | ···················· | ···················· | ···················· |
| L | 3806 hCoV-19/England/ALDP-9EA23A/2020\|EPI_ISL_580066\|20 | ···················· | ···················· | ····················· | ···················· | ···················· | ···················· |
| L | 3807 hCoV-19/England/ALDP-9EA20D/2020\|EPI_ISL_580067\|20 | ···················· | ···················· | ····················· | ···················· | ···················· | ···················· |
| L | 3808 hCoV-19/England/ALDP-9EA58C/2020\|EPI_ISL_580068\|20 | ···················· | ···················· | ····················· | ···················· | ···················· | ···················· |
| L | 3809 hCoV-19/England/ALDP-9EA7FF/2020\|EPI_ISL_580069\|20 | ···················· | ···················· | ····················· | ···················· | ···················· | ···················· |
| L | 3810 hCoV-19/England/ALDP-9EAA05/2020\|EPI_ISL_580070\|20 | ···················· | ···················· | ····················· | ···················· | ···················· | ···················· |
| L | 3811 hCoV-19/England/ALDP-9EAF60/2020\|EPI_ISL_580071\|20 | ···················· | ···················· | ····················· | ···················· | ···················· | ···················· |
| L | 3812 hCoV-19/England/ALDP-9EAAAB/2020\|EPI_ISL_580072\|20 | ···················· | ···················· | ····················· | ···················· | ···················· | ···················· |
| L | 3813 hCoV-19/England/ALDP-9EA9DB/2020\|EPI_ISL_580074\|20 | ···················· | ···················· | ····················· | ···················· | ···················· | ···················· |
| L | 3814 hCoV-19/England/ALDP-9EA80B/2020\|EPI_ISL_580076\|20 | ···················· | ·A·················· | ····················· | ···················· | ···················· | ···················· |
| L | 3815 hCoV-19/England/ALDP-9EA48F/2020\|EPI_ISL_580077\|20 | ···················· | ···················· | ····················· | ···················· | ···················· | ···················· |
| L | 3816 hCoV-19/England/ALDP-9EA72C/2020\|EPI_ISL_580078\|20 | ···················· | ···················· | ····················· | ···················· | ···················· | ···················· |
| L | 3817 hCoV-19/England/ALDP-9EB08A/2020\|EPI_ISL_580079\|20 | ···················· | ···················· | ····················· | ···················· | ···················· | ···················· |
| L | 3818 hCoV-19/England/ALDP-9EAF06/2020\|EPI_ISL_580080\|20 | ···················· | ···················· | ····················· | ···················· | ···················· | ···················· |
| L | 3819 hCoV-19/England/ALDP-9E76B9/2020\|EPI_ISL_580081\|20 | ···················· | ···················· | ····················· | ···················· | ···················· | ···················· |
| L | 3820 hCoV-19/England/ALDP-9EA917/2020\|EPI_ISL_580082\|20 | ···················· | ·A·················· | ····················· | ···················· | ···················· | ···A················ |
| L | 3821 hCoV-19/England/ALDP-9EA4CB/2020\|EPI_ISL_580083\|20 | ···················· | ···················· | ····················· | ···················· | ···················· | ···················· |
| L | 3822 hCoV-19/England/ALDP-9E9CF1/2020\|EPI_ISL_580084\|20 | ···················· | ···················· | ····················· | ···················· | ···················· | ···················· |
| L | 3823 hCoV-19/England/ALDP-9EA847/2020\|EPI_ISL_580085\|20 | ···················· | ···················· | ····················· | ···················· | ···················· | ···················· |
| L | 3824 hCoV-19/England/ALDP-9EA865/2020\|EPI_ISL_580086\|20 | ···················· | ···················· | ····················· | ···················· | ···················· | ···················· |
| L | 3825 hCoV-19/England/MILK-9D4A3B/2020\|EPI_ISL_581374\|20 | ···················· | ···················· | ····················· | ···················· | ···················· | ···················· |
| L | 3826 hCoV-19/England/MILK-9C44A9/2020\|EPI_ISL_581376\|20 | ···················· | ···················· | ····················· | ···················· | ···················· | ···················· |

| | | | | | | | |
|---|---|---|---|---|---|---|---|
| L | 3827 hCoV-19/England/QEUH-9C85FD/2020\|EPI_ISL_581377\|20 | ···················· | ···················· | ···················· | ···················· | ···················· | ···················· |
| L | 3828 hCoV-19/England/ALDP-9C4CB0/2020\|EPI_ISL_581378\|20 | ···················· | ···················· | ···················· | ···················· | ···················· | ···················· |
| L | 3829 hCoV-19/England/ALDP-9C5790/2020\|EPI_ISL_581379\|20 | ···················· | ···················· | ···················· | ···················· | ···················· | ···················· |
| L | 3830 hCoV-19/England/QEUH-9B8420/2020\|EPI_ISL_581381\|20 | ···················· | ···················· | ···················· | ···················· | ···················· | ···················· |
| L | 3831 hCoV-19/England/QEUH-998857/2020\|EPI_ISL_581384\|20 | ···················· | ·A·················· | ···················· | ···················· | ···················· | ··A················· |
| L | 3832 hCoV-19/England/MILK-99C49B/2020\|EPI_ISL_581385\|20 | ···················· | ···················· | ···················· | ···················· | ·····T·············· | ···················· |
| L | 3833 hCoV-19/England/MILK-99DE9C/2020\|EPI_ISL_581386\|20 | ···················· | ···················· | ···················· | ···················· | ···················· | ···················· |
| L | 3834 hCoV-19/England/QEUH-9992FE/2020\|EPI_ISL_581387\|20 | ···················· | ·A·················· | ···················· | ···················· | ···················· | ··A················· |
| L | 3835 hCoV-19/England/MILK-97786D/2020\|EPI_ISL_581389\|20 | ···················· | ···················· | ···················· | ···················· | ···················· | ···················· |
| L | 3836 hCoV-19/England/QEUH-98F325/2020\|EPI_ISL_581395\|20 | ···················· | ···················· | ···················· | ···················· | ···················· | ···················· |
| L | 3837 hCoV-19/England/CAMC-991135/2020\|EPI_ISL_581396\|20 | ···················· | ···················· | ···················· | ···················· | ···················· | ···················· |
| L | 3838 hCoV-19/England/MILK-975805/2020\|EPI_ISL_581397\|20 | ···················· | ···················· | ···················· | ···················· | ···················· | ···················· |
| L | 3839 hCoV-19/England/MILK-970451/2020\|EPI_ISL_581398\|20 | ···················· | ·A·················· | ···················· | ···················· | ···················· | ··A················· |
| L | 3840 hCoV-19/England/MILK-970CB3/2020\|EPI_ISL_581399\|20 | ···················· | ···················· | ···················· | ···················· | ···················· | ···················· |
| L | 3841 hCoV-19/USA/MI-QDX-1120/2020\|EPI_ISL_571850\|2020-0 | ···················· | ···················· | ···················· | ···················· | ···················· | ···················· |
| L | 3842 hCoV-19/USA/MI-QDX-1121/2020\|EPI_ISL_571851\|2020-0 | ···················· | ···················· | ···················· | ···················· | ···················· | ···················· |
| L | 3843 hCoV-19/USA/MI-QDX-1122/2020\|EPI_ISL_571852\|2020-0 | ···················· | ···················· | ···················· | ···················· | ···················· | ···················· |
| L | 3844 hCoV-19/USA/MI-QDX-1123/2020\|EPI_ISL_571853\|2020-0 | ···················· | ···················· | ···················· | ···················· | ···················· | ···················· |
| L | 3845 hCoV-19/USA/MI-QDX-1124/2020\|EPI_ISL_571854\|2020-0 | ···················· | ···················· | ···················· | ···················· | ···················· | ···················· |
| L | 3846 hCoV-19/USA/MI-QDX-1125/2020\|EPI_ISL_571855\|2020-0 | ···················· | ···················· | ···················· | ···················· | ···················· | ···················· |
| L | 3847 hCoV-19/USA/MI-QDX-1126/2020\|EPI_ISL_571856\|2020-0 | ···················· | ···················· | ···················· | ···················· | ···················· | ···················· |
| L | 3848 hCoV-19/USA/MI-QDX-1127/2020\|EPI_ISL_571857\|2020-0 | ···················· | ···················· | ···················· | ···················· | ···················· | ···················· |
| L | 3849 hCoV-19/USA/MI-QDX-1128/2020\|EPI_ISL_571858\|2020-0 | ···················· | ···················· | ···················· | ···················· | ···················· | ···················· |
| L | 3850 hCoV-19/USA/MI-QDX-1130/2020\|EPI_ISL_571860\|2020-0 | ···················· | ···················· | ···················· | ···················· | ···················· | ···················· |
| L | 3851 hCoV-19/USA/MI-QDX-1132/2020\|EPI_ISL_571862\|2020-0 | ···················· | ···················· | ···················· | ···················· | ···················· | ···················· |
| L | 3852 hCoV-19/USA/MI-QDX-1133/2020\|EPI_ISL_571863\|2020-0 | ···················· | ···················· | ···················· | ···················· | ···················· | ···················· |
| L | 3853 hCoV-19/USA/MI-QDX-1134/2020\|EPI_ISL_571864\|2020-0 | ···················· | ···················· | ···················· | ···················· | ···················· | ···················· |
| L | 3854 hCoV-19/USA/MI-QDX-1135/2020\|EPI_ISL_571865\|2020-0 | ···················· | ···················· | ···················· | ···················· | ···················· | ···················· |
| L | 3855 hCoV-19/USA/MI-QDX-1136/2020\|EPI_ISL_571866\|2020-0 | ···················· | ···················· | ···················· | ···················· | ···················· | ···················· |
| L | 3856 hCoV-19/USA/MI-QDX-1137/2020\|EPI_ISL_571867\|2020-0 | ···················· | ···················· | ···················· | ···················· | ···················· | ···················· |
| L | 3857 hCoV-19/USA/MI-QDX-1138/2020\|EPI_ISL_571868\|2020-0 | ···················· | ···················· | ···················· | ···················· | ···················· | ···················· |
| L | 3858 hCoV-19/USA/MI-QDX-1139/2020\|EPI_ISL_571869\|2020-0 | ···················· | ···················· | ···················· | ···················· | ···················· | ···················· |
| L | 3859 hCoV-19/USA/MI-QDX-1140/2020\|EPI_ISL_571870\|2020-0 | ···················· | ···················· | ···················· | ···················· | ···················· | ···················· |
| L | 3860 hCoV-19/USA/MI-QDX-1141/2020\|EPI_ISL_571871\|2020-0 | ···················· | ···················· | ···················· | ···················· | ···················· | ···················· |
| L | 3861 hCoV-19/USA/MI-QDX-1142/2020\|EPI_ISL_571872\|2020-0 | ···················· | ···················· | ···················· | ···················· | ···················· | ···················· |
| L | 3862 hCoV-19/USA/MI-QDX-1143/2020\|EPI_ISL_571873\|2020-0 | ···················· | ···················· | ···················· | ···················· | ···················· | ···················· |
| L | 3863 hCoV-19/USA/MI-QDX-1144/2020\|EPI_ISL_571874\|2020-0 | ···················· | ···················· | ···················· | ···················· | ···················· | ···················· |
| L | 3864 hCoV-19/USA/MI-QDX-1145/2020\|EPI_ISL_571875\|2020-0 | ···················· | ···················· | ···················· | ···················· | ···················· | ···················· |
| L | 3865 hCoV-19/USA/MI-QDX-1146/2020\|EPI_ISL_571876\|2020-0 | ···················· | ···················· | ···················· | ···················· | ···················· | ···················· |
| L | 3866 hCoV-19/USA/MI-QDX-1147/2020\|EPI_ISL_571877\|2020-0 | ···················· | ···················· | ···················· | ···················· | ···················· | ···················· |
| L | 3867 hCoV-19/USA/MI-QDX-1148/2020\|EPI_ISL_571878\|2020-0 | ···················· | ···················· | ···················· | ···················· | ···················· | ···················· |
| L | 3868 hCoV-19/USA/MI-QDX-1149/2020\|EPI_ISL_571879\|2020-0 | ···················· | ···················· | ···················· | ···················· | ···················· | ···················· |
| L | 3869 hCoV-19/USA/MI-QDX-1150/2020\|EPI_ISL_571880\|2020-0 | ···················· | ···················· | ···················· | ···················· | ···················· | ···················· |
| L | 3870 hCoV-19/USA/MI-QDX-1151/2020\|EPI_ISL_571881\|2020-0 | ···················· | ···················· | ···················· | ···················· | ···················· | ···················· |
| L | 3871 hCoV-19/USA/MI-QDX-1152/2020\|EPI_ISL_571882\|2020-0 | ···················· | ···················· | ···················· | ···················· | ···················· | ···················· |
| L | 3872 hCoV-19/USA/MI-QDX-1153/2020\|EPI_ISL_571883\|2020-0 | ···················· | ···················· | ···················· | ···················· | ···················· | ···················· |
| L | 3873 hCoV-19/USA/MI-QDX-1154/2020\|EPI_ISL_571884\|2020-0 | ···················· | ···················· | ···················· | ···················· | ···················· | ···················· |
| L | 3874 hCoV-19/USA/MI-QDX-1155/2020\|EPI_ISL_571885\|2020-0 | ···················· | ···················· | ···················· | ···················· | ···················· | ···················· |
| L | 3875 hCoV-19/USA/MI-QDX-1156/2020\|EPI_ISL_571886\|2020-0 | ···················· | ···················· | ···················· | ···················· | ···················· | ···················· |
| L | 3876 hCoV-19/USA/MI-QDX-1157/2020\|EPI_ISL_571887\|2020-0 | ···················· | ···················· | ···················· | ···················· | ···················· | ···················· |
| L | 3877 hCoV-19/USA/MI-QDX-1158/2020\|EPI_ISL_571888\|2020-0 | ···················· | ···················· | ···················· | ···················· | ···················· | ···················· |
| L | 3878 hCoV-19/USA/MI-QDX-1159/2020\|EPI_ISL_571889\|2020-0 | ···················· | ···················· | ···················· | ···················· | ···················· | ···················· |
| L | 3879 hCoV-19/USA/MI-QDX-1160/2020\|EPI_ISL_571890\|2020-0 | ···················· | ···················· | ···················· | ···················· | ···················· | ···················· |
| L | 3880 hCoV-19/USA/MI-QDX-1161/2020\|EPI_ISL_571891\|2020-0 | ···················· | ···················· | ···················· | ···················· | ···················· | ···················· |
| L | 3881 hCoV-19/USA/MI-QDX-1162/2020\|EPI_ISL_571892\|2020-0 | ···················· | ···················· | ···················· | ···················· | ···················· | ···················· |
| L | 3882 hCoV-19/USA/MI-QDX-1163/2020\|EPI_ISL_571893\|2020-0 | ···················· | ···················· | ···················· | ···················· | ···················· | ···················· |
| L | 3883 hCoV-19/USA/MI-QDX-1164/2020\|EPI_ISL_571894\|2020-0 | ···················· | ···················· | ···················· | ···················· | ···················· | ···················· |
| L | 3884 hCoV-19/USA/MI-QDX-1165/2020\|EPI_ISL_571895\|2020-0 | ···················· | ···················· | ···················· | ···················· | ···················· | ···················· |
| L | 3885 hCoV-19/USA/MI-QDX-1167/2020\|EPI_ISL_571897\|2020-0 | ···················· | ···················· | ···················· | ···················· | ···················· | ···················· |
| L | 3886 hCoV-19/USA/MS-QDX-1071/2020\|EPI_ISL_571898\|2020-0 | ···················· | ···················· | ···················· | ···················· | ···················· | ···················· |
| L | 3887 hCoV-19/USA/PR-QDX-1074/2020\|EPI_ISL_571899\|2020-0 | ···················· | ···················· | ···················· | ···················· | ···················· | ···················· |
| L | 3888 hCoV-19/Wales/PHWC-16C3DB/2020\|EPI_ISL_574051\|2020 | ···················· | ···················· | ···················· | ···················· | ···················· | ···················· |
| L | 3889 hCoV-19/Wales/PHWC-16C3EA/2020\|EPI_ISL_574052\|2020 | ···················· | ···················· | ···················· | ···················· | ···················· | ···················· |
| L | 3890 hCoV-19/Wales/PHWC-16C405/2020\|EPI_ISL_574053\|2020 | ···················· | ···················· | ···················· | ···················· | ···················· | ···················· |
| L | 3891 hCoV-19/Wales/PHWC-16C414/2020\|EPI_ISL_574054\|2020 | ···················· | ···················· | ···················· | ···················· | ···················· | ···················· |
| L | 3892 hCoV-19/Wales/PHWC-16C450/2020\|EPI_ISL_574057\|2020 | ···················· | ···················· | ···················· | ···················· | ···················· | ···················· |
| L | 3893 hCoV-19/Wales/PHWC-16C4BA/2020\|EPI_ISL_574059\|2020 | ···················· | ···················· | ···················· | ···················· | ···················· | ···················· |
| L | 3894 hCoV-19/Wales/PHWC-16C4C9/2020\|EPI_ISL_574060\|2020 | ···················· | ···················· | ···················· | ···················· | ···················· | ···················· |
| L | 3895 hCoV-19/Wales/PHWC-16C4E7/2020\|EPI_ISL_574061\|2020 | ···················· | ···················· | ···················· | ···················· | ···················· | ···················· |
| L | 3896 hCoV-19/Wales/PHWC-16C4F6/2020\|EPI_ISL_574062\|2020 | ···················· | ···················· | ···················· | ···················· | ···················· | ···················· |
| L | 3897 hCoV-19/Wales/PHWC-16C502/2020\|EPI_ISL_574063\|2020 | ···················· | ···················· | ···················· | ···················· | ···················· | ···················· |
| L | 3898 hCoV-19/Wales/PHWC-16C53F/2020\|EPI_ISL_574064\|2020 | ···················· | ···················· | ···················· | ···················· | ···················· | ···················· |
| L | 3899 hCoV-19/Wales/PHWC-16C56C/2020\|EPI_ISL_574065\|2020 | ···················· | ···················· | ···················· | ···················· | ···················· | ···················· |
| L | 3900 hCoV-19/Wales/PHWC-16C57B/2020\|EPI_ISL_574066\|2020 | ···················· | ···················· | ···················· | ···················· | ···················· | ···················· |
| L | 3902 hCoV-19/Wales/PHWC-16C599/2020\|EPI_ISL_574068\|2020 | ···················· | ···················· | ···················· | ···················· | ···················· | ···················· |
| L | 3905 hCoV-19/Wales/PHWC-16D0E2/2020\|EPI_ISL_574201\|2020 | ···················· | ···················· | ···················· | ···················· | ···················· | ···················· |
| L | 3906 hCoV-19/Scotland/QEUH-9DA859/2020\|EPI_ISL_575382\|2 | ···················· | ···················· | ···················· | ···················· | ···················· | ···················· |
| L | 3907 hCoV-19/Scotland/QEUH-9DBF80/2020\|EPI_ISL_575383\|2 | ···················· | ···················· | ···················· | ···················· | ···················· | ···················· |
| L | 3908 hCoV-19/Scotland/QEUH-9DAA9F/2020\|EPI_ISL_575384\|2 | ···················· | ···················· | ···················· | ···················· | ···················· | ···················· |
| L | 3909 hCoV-19/Scotland/QEUH-9DA80E/2020\|EPI_ISL_575385\|2 | ···················· | ···················· | ···················· | ···················· | ···················· | ···················· |

| | | | | | | | | |
|---|---|---|---|---|---|---|---|---|
| L | 3910 hCoV-19/Scotland/QEUH-9DA84A/2020\|EPI_ISL_575390\|2 | ···················· | ···················· | ···················· | ···················· | ···················· | ···················· | |
| L | 3911 hCoV-19/Scotland/QEUH-9DAA80/2020\|EPI_ISL_575391\|2 | ···················· | ···················· | ···················· | ···················· | ···················· | ···················· | |
| L | 3912 hCoV-19/Scotland/QEUH-9DBFDB/2020\|EPI_ISL_575392\|2 | ···················· | ···················· | ···················· | ···················· | ···················· | ···················· | |
| L | 3913 hCoV-19/Scotland/QEUH-9DA7D4/2020\|EPI_ISL_575393\|2 | ···················· | ···················· | ···················· | ···················· | ···················· | ···················· | |
| L | 3914 hCoV-19/Scotland/QEUH-9DAABD/2020\|EPI_ISL_575394\|2 | ···················· | ···················· | ···················· | ···················· | ···················· | ···················· | |
| L | 3915 hCoV-19/Scotland/QEUH-9DC1F2/2020\|EPI_ISL_575395\|2 | ···················· | ···················· | ···················· | ···················· | ···················· | ···················· | |
| L | 3916 hCoV-19/Scotland/QEUH-9DC0D7/2020\|EPI_ISL_575396\|2 | ···················· | ···················· | ···················· | ···················· | ···················· | ···················· | |
| L | 3917 hCoV-19/Scotland/QEUH-9DA7B6/2020\|EPI_ISL_575397\|2 | ···················· | ···················· | ···················· | ···················· | ···················· | ···················· | |
| L | 3918 hCoV-19/Scotland/QEUH-9DA5E9/2020\|EPI_ISL_575398\|2 | ···················· | ···················· | ···················· | ···················· | ···················· | ···················· | |
| L | 3919 hCoV-19/England/ALDP-9CAE87/2020\|EPI_ISL_575399\|20 | ···················· | ···················· | ···················· | ···················· | ···················· | ···················· | |
| L | 3920 hCoV-19/England/ALDP-9CB84C/2020\|EPI_ISL_575500\|20 | ···················· | ···················· | ···················· | ···················· | ···T················ | ···················· | |
| L | 3921 hCoV-19/England/ALDP-9CB7C7/2020\|EPI_ISL_575501\|20 | ···················· | ···················· | ···················· | ···················· | ···················· | ···················· | |
| L | 3922 hCoV-19/England/ALDP-9CBB34/2020\|EPI_ISL_575502\|20 | ···················· | ···················· | ···················· | ···················· | ···················· | ···················· | |
| L | 3923 hCoV-19/England/ALDP-9CB123/2020\|EPI_ISL_575503\|20 | ···················· | ···················· | ···················· | ···················· | ···················· | ···················· | |
| L | 3924 hCoV-19/England/ALDP-9CAC8D/2020\|EPI_ISL_575505\|20 | ···················· | ···················· | ···················· | ···················· | ···················· | ···················· | |
| L | 3925 hCoV-19/England/ALDP-9CAD4E/2020\|EPI_ISL_575506\|20 | ···················· | ···················· | ···················· | ···················· | ···················· | ···················· | |
| L | 3926 hCoV-19/England/ALDP-9CB8F1/2020\|EPI_ISL_575507\|20 | ···················· | ···················· | ···················· | ···················· | ···················· | ···················· | |
| L | 3927 hCoV-19/England/ALDP-9CBEE0/2020\|EPI_ISL_575508\|20 | ···················· | ···················· | ···················· | ···················· | ···················· | ···················· | |
| L | 3928 hCoV-19/England/ALDP-9CB4FD/2020\|EPI_ISL_575509\|20 | ···················· | ···················· | ···················· | ···················· | ···················· | ···················· | |
| L | 3929 hCoV-19/England/ALDP-9CAF84/2020\|EPI_ISL_575510\|20 | ···················· | ···················· | ···················· | ···················· | ···················· | ···················· | |
| L | 3930 hCoV-19/England/ALDP-9CBD3E/2020\|EPI_ISL_575511\|20 | ···················· | ···················· | ···················· | ···················· | ···················· | ···················· | |
| L | 3931 hCoV-19/England/ALDP-9CACAB/2020\|EPI_ISL_575512\|20 | ···················· | ···················· | ···················· | ···················· | ···················· | ···················· | |
| L | 3932 hCoV-19/England/ALDP-9CAEA5/2020\|EPI_ISL_575513\|20 | ···················· | ···················· | ···················· | ···················· | ···················· | ···················· | |
| L | 3933 hCoV-19/England/ALDP-9CB527/2020\|EPI_ISL_575514\|20 | ···················· | ···················· | ···················· | ···················· | ···················· | ···················· | |
| L | 3934 hCoV-19/England/ALDP-9CBD4D/2020\|EPI_ISL_575515\|20 | ···················· | ···················· | ···················· | ···················· | ···················· | ···················· | |
| L | 3935 hCoV-19/England/ALDP-9CABBD/2020\|EPI_ISL_575516\|20 | ···················· | ···················· | ···················· | ···················· | ···················· | ···················· | |
| L | 3936 hCoV-19/England/ALDP-9CB2B7/2020\|EPI_ISL_575517\|20 | ···················· | ·A·················· | ···················· | ···················· | ···················· | ··A················· | |
| L | 3937 hCoV-19/England/ALDP-9CB30F/2020\|EPI_ISL_575518\|20 | ···················· | ···················· | ···················· | ···················· | ···················· | ···················· | |
| L | 3938 hCoV-19/England/ALDP-9CBB52/2020\|EPI_ISL_575519\|20 | ···················· | ···················· | ···················· | ···················· | ···················· | ···················· | |
| L | 3939 hCoV-19/England/ALDP-9CB712/2020\|EPI_ISL_575520\|20 | ···················· | ···················· | ···················· | ···················· | ···················· | ···················· | |
| L | 3940 hCoV-19/England/ALDP-9CB800/2020\|EPI_ISL_575522\|20 | ···················· | ···················· | ···················· | ···················· | ···················· | ···················· | |
| L | 3941 hCoV-19/England/ALDP-9CABAE/2020\|EPI_ISL_575523\|20 | ···················· | ·A·················· | ···················· | ···················· | ···················· | ··A················· | |
| L | 3942 hCoV-19/England/ALDP-9CC0BC/2020\|EPI_ISL_575524\|20 | ···················· | ···················· | ···················· | ···················· | ···················· | ···················· | |
| L | 3943 hCoV-19/England/ALDP-9CBEFF/2020\|EPI_ISL_575525\|20 | ···················· | ···················· | ···················· | ···················· | ···················· | ···················· | |
| L | 3944 hCoV-19/England/ALDP-9CBAA0/2020\|EPI_ISL_575526\|20 | ···················· | ·A·················· | ···················· | ···················· | ···················· | ···················· | |
| L | 3945 hCoV-19/England/ALDP-9CB7A9/2020\|EPI_ISL_575527\|20 | ···················· | ···················· | ···················· | ···················· | ···················· | ···················· | |
| L | 3946 hCoV-19/England/ALDP-9CACE7/2020\|EPI_ISL_575528\|20 | ···················· | ···················· | ···················· | ···················· | ···················· | ···················· | |
| L | 3947 hCoV-19/England/ALDP-9CBB07/2020\|EPI_ISL_575529\|20 | ···················· | ···················· | ···················· | ···················· | ···················· | ···················· | |
| L | 3948 hCoV-19/England/ALDP-9CB475/2020\|EPI_ISL_575530\|20 | ···················· | ···················· | ···················· | ···················· | ···················· | ···················· | |
| L | 3949 hCoV-19/England/ALDP-9CA9FF/2020\|EPI_ISL_575531\|20 | ···················· | ·A·················· | ···················· | ···················· | ···················· | ··A················· | |
| L | 3950 hCoV-19/England/ALDP-9CBF38/2020\|EPI_ISL_575532\|20 | ···················· | ···················· | ···················· | ···················· | ···················· | ···················· | |
| L | 3951 hCoV-19/England/ALDP-9D9BD9/2020\|EPI_ISL_576800\|20 | ···················· | ···················· | ···················· | ···················· | ···················· | ···················· | |
| L | 3952 hCoV-19/England/ALDP-9D9BF7/2020\|EPI_ISL_576802\|20 | ···················· | ···················· | ···················· | ···················· | ···················· | ···················· | |
| L | 3953 hCoV-19/England/ALDP-9D9C03/2020\|EPI_ISL_576803\|20 | ···················· | ···················· | ···················· | ···················· | ···················· | ···················· | |
| L | 3954 hCoV-19/England/ALDP-9D9C12/2020\|EPI_ISL_576804\|20 | ···················· | ···················· | ···················· | ···················· | ···················· | ···················· | |
| L | 3955 hCoV-19/England/ALDP-9D9C21/2020\|EPI_ISL_576805\|20 | ···················· | ···················· | ···················· | ···················· | ···················· | ···················· | |
| L | 3956 hCoV-19/England/ALDP-9D9E0D/2020\|EPI_ISL_576820\|20 | ···················· | ···················· | ···················· | ···················· | ···················· | ···················· | |
| L | 3957 hCoV-19/England/ALDP-9D9E3A/2020\|EPI_ISL_576821\|20 | ···················· | ···················· | ···················· | ···················· | ···················· | ···················· | |
| L | 3958 hCoV-19/England/ALDP-9D9E49/2020\|EPI_ISL_576822\|20 | ···················· | ·····A·············· | ···················· | ···················· | ···················· | ···················· | |
| L | 3959 hCoV-19/England/ALDP-9D9EA3/2020\|EPI_ISL_576823\|20 | ···················· | ···················· | ···················· | ···················· | ···················· | ···················· | |
| L | 3960 hCoV-19/England/ALDP-9D9EEF/2020\|EPI_ISL_576824\|20 | ···················· | ···················· | ···················· | ···················· | ···················· | ···················· | |
| L | 3961 hCoV-19/England/ALDP-9D9EFE/2020\|EPI_ISL_576825\|20 | ···················· | ···················· | ···················· | ···················· | ···················· | ···················· | |
| L | 3962 hCoV-19/England/ALDP-9D9F0A/2020\|EPI_ISL_576826\|20 | ···················· | ···················· | ···················· | ···················· | ···················· | ···················· | |
| L | 3963 hCoV-19/England/ALDP-9D9F19/2020\|EPI_ISL_576827\|20 | ···················· | ···················· | ···················· | ···················· | ···················· | ···················· | |
| L | 3964 hCoV-19/England/ALDP-9D9F46/2020\|EPI_ISL_576828\|20 | ···················· | ···················· | ···················· | ···················· | ···················· | ···················· | |
| L | 3965 hCoV-19/England/ALDP-9D9F73/2020\|EPI_ISL_576829\|20 | ···················· | ···················· | ···················· | ···················· | ···················· | ···················· | |
| L | 3966 hCoV-19/England/ALDP-9D9F82/2020\|EPI_ISL_576830\|20 | ···················· | ···················· | ···················· | ···················· | ···················· | ···················· | |
| L | 3967 hCoV-19/England/ALDP-9D9FA0/2020\|EPI_ISL_576831\|20 | ···················· | ···················· | ···················· | ···················· | ·····T·············· | ···················· | |
| L | 3968 hCoV-19/England/ALDP-9D9FBF/2020\|EPI_ISL_576832\|20 | ···················· | ···················· | ···················· | ···················· | ·····T·············· | ···················· | |
| L | 3969 hCoV-19/England/ALDP-9D9FCE/2020\|EPI_ISL_576833\|20 | ···················· | ···················· | ···················· | ···················· | ···················· | ···················· | |
| L | 3970 hCoV-19/England/ALDP-9D9FDD/2020\|EPI_ISL_576834\|20 | ···················· | ···················· | ···················· | ···················· | ···················· | ···················· | |
| L | 3971 hCoV-19/England/ALDP-9D9FFB/2020\|EPI_ISL_576835\|20 | ···················· | ···················· | ···················· | ···················· | ···················· | ···················· | |
| L | 3972 hCoV-19/England/ALDP-9DA015/2020\|EPI_ISL_576836\|20 | ···················· | ···················· | ···················· | ···················· | ···················· | ···················· | |
| L | 3973 hCoV-19/England/ALDP-9DA024/2020\|EPI_ISL_576837\|20 | ···················· | ···················· | ···················· | ···················· | ···················· | ···················· | |
| L | 3974 hCoV-19/England/ALDP-9DA033/2020\|EPI_ISL_576838\|20 | ···················· | ···················· | ···················· | ···················· | ···················· | ···················· | |
| L | 3975 hCoV-19/England/ALDP-9DA042/2020\|EPI_ISL_576839\|20 | ···················· | ···················· | ···················· | ···················· | ···················· | ···················· | |
| L | 3976 hCoV-19/England/ALDP-9DA060/2020\|EPI_ISL_576840\|20 | ···················· | ···················· | ···················· | ···················· | ···················· | ···················· | |
| L | 3977 hCoV-19/England/ALDP-9DA07F/2020\|EPI_ISL_576841\|20 | ···················· | ···················· | ···················· | ···················· | ···················· | ···················· | |
| L | 3978 hCoV-19/England/ALDP-9DA08E/2020\|EPI_ISL_576842\|20 | ···················· | ···················· | ···················· | ···················· | ···················· | ···················· | |
| L | 3979 hCoV-19/England/ALDP-9DA0BB/2020\|EPI_ISL_576843\|20 | ···················· | ···················· | ···················· | ···················· | ···················· | ···················· | |
| L | 3980 hCoV-19/England/ALDP-9DA0D9/2020\|EPI_ISL_576844\|20 | ···················· | ···················· | ···················· | ···················· | ···T················ | ···················· | |
| L | 3981 hCoV-19/England/ALDP-9DA0E8/2020\|EPI_ISL_576845\|20 | ···················· | ···················· | ···················· | ···················· | ···················· | ···················· | |
| L | 3982 hCoV-19/England/ALDP-9DA0F7/2020\|EPI_ISL_576846\|20 | ···················· | ···················· | ···················· | ···················· | ···················· | ···················· | |
| L | 3983 hCoV-19/England/ALDP-9DA103/2020\|EPI_ISL_576847\|20 | ···················· | ···················· | ···················· | ···················· | ···················· | ···················· | |
| L | 3984 hCoV-19/England/ALDP-9DA112/2020\|EPI_ISL_576848\|20 | ···················· | ···················· | ···················· | ···················· | ···················· | ···················· | |
| L | 3985 hCoV-19/England/ALDP-9DA121/2020\|EPI_ISL_576849\|20 | ···················· | ···················· | ···················· | ···················· | ···················· | ···················· | |
| L | 3986 hCoV-19/England/ALDP-9DA130/2020\|EPI_ISL_576850\|20 | ···················· | ··---··············· | ···················· | ···················· | ···················· | ···················· | 1 |
| L | 3987 hCoV-19/England/ALDP-9DA14F/2020\|EPI_ISL_576851\|20 | ···················· | ···················· | ···················· | ···················· | ···················· | ···················· | |
| L | 3988 hCoV-19/England/ALDP-9DA16D/2020\|EPI_ISL_576853\|20 | ···················· | ···················· | ···················· | ···················· | ···················· | ···················· | |
| L | 3989 hCoV-19/England/ALDP-9DA17C/2020\|EPI_ISL_576854\|20 | ···················· | ···················· | ···················· | ···················· | ···················· | ···················· | |

L 3990 hCoV-19/England/ALDP-9DA18B/2020|EPI_ISL_576855|20 ···················· ···················· ····················· ···················· ···················· ····················
L 3991 hCoV-19/England/ALDP-9DA22E/2020|EPI_ISL_576857|20 ···················· ···················· ····················· ···················· ···················· ····················
L 3992 hCoV-19/England/ALDP-9DA24C/2020|EPI_ISL_576858|20 ···················· ···················· ····················· ···················· ···················· ····················
L 3993 hCoV-19/England/ALDP-9DA25B/2020|EPI_ISL_576859|20 ···················· ···················· ····················· ···················· ···················· ····················
L 3994 hCoV-19/England/ALDP-9DA2D3/2020|EPI_ISL_576861|20 ···················· ···················· ····················· ···················· ···················· ····················
L 3995 hCoV-19/England/ALDP-9DA31C/2020|EPI_ISL_576863|20 ···················· ···················· ····················· ···················· ···················· ····················
L 3996 hCoV-19/England/ALDP-9E9C1F/2020|EPI_ISL_580060|20 ···················· ·A·················· ····················· ···················· ···················· ··A·················
L 3997 hCoV-19/England/ALDP-9EA267/2020|EPI_ISL_580061|20 ···················· ···················· ····················· ···················· ···················· ····················
L 3998 hCoV-19/England/ALDP-9EAA8D/2020|EPI_ISL_580064|20 ···················· ···················· ····················· ···················· ···················· ····················
L 3999 hCoV-19/England/ALDP-9EA935/2020|EPI_ISL_580088|20 ···················· ···················· ····················· ···················· ···················· ····················
L 4000 hCoV-19/England/ALDP-9EA953/2020|EPI_ISL_580089|20 ···················· ···················· ····················· ···················· ···················· ····················
L 4001 hCoV-19/England/ALDP-9EAC0F/2020|EPI_ISL_580090|20 ···················· ···················· ····················· ···················· ···················· ····················
L 4002 hCoV-19/England/ALDP-9EA70E/2020|EPI_ISL_580091|20 ···················· ···················· ····················· ···················· ···················· ····················
L 4003 hCoV-19/England/ALDP-9EAC2D/2020|EPI_ISL_580092|20 ···················· ···················· ····················· ···················· ···················· ····················
L 4004 hCoV-19/England/ALDP-9E9C5B/2020|EPI_ISL_580093|20 ···················· ···················· ····················· ···················· ···················· ····················
L 4005 hCoV-19/England/ALDP-9EA5AA/2020|EPI_ISL_580094|20 ···················· ···················· ····················· ···················· ···················· ····················
L 4006 hCoV-19/England/ALDP-9EA021/2020|EPI_ISL_580095|20 ···················· ···················· ····················· ···················· ···················· ····················
L 4007 hCoV-19/England/ALDP-9E9F61/2020|EPI_ISL_580096|20 ···················· ···················· ····················· ···················· ···················· ····················
L 4008 hCoV-19/England/ALDP-9E9FCB/2020|EPI_ISL_580098|20 ···················· ···················· ····················· ···················· ···················· ····················
L 4009 hCoV-19/England/ALDP-9EA504/2020|EPI_ISL_580099|20 ···················· ···················· ····················· ···················· ···················· ····················
L 4010 hCoV-19/England/MILK-9E2D31/2020|EPI_ISL_581200|20 ···················· ···················· ····················· ···················· ···················· ····················
L 4011 hCoV-19/Wales/MILK-9E2162/2020|EPI_ISL_581201|2020 ···················· ···················· ····················· ···················· ···················· ····················
L 4012 hCoV-19/Wales/MILK-9E2A94/2020|EPI_ISL_581202|2020 ···················· ···················· ····················· ···················· ···················· ····················
L 4013 hCoV-19/England/MILK-9E2A67/2020|EPI_ISL_581203|20 ···················· ···················· ····················· ···················· ···················· ····················
L 4014 hCoV-19/England/MILK-9E1DD8/2020|EPI_ISL_581205|20 ···················· ···················· ····················· ···················· ···················· ····················
L 4015 hCoV-19/England/MILK-9E2E7A/2020|EPI_ISL_581206|20 ···················· ·A·················· ····················· ···················· ···················· ··A·················
L 4016 hCoV-19/England/MILK-9E1F1E/2020|EPI_ISL_581207|20 ···················· ···················· ····················· ···················· ···················· ····················
L 4017 hCoV-19/England/MILK-9E1C80/2020|EPI_ISL_581208|20 ···················· ·A·················· ····················· ···················· ···················· ····················
L 4018 hCoV-19/Wales/MILK-9E326E/2020|EPI_ISL_581209|2020 ···················· ···················· ····················· ···················· ···················· ····················
L 4019 hCoV-19/England/MILK-9E2AFE/2020|EPI_ISL_581210|20 ···················· ···················· ····················· ···················· ···················· ····················
L 4020 hCoV-19/England/MILK-9E2A49/2020|EPI_ISL_581211|20 ···················· ···················· ····················· ···················· ···················· ····················
L 4021 hCoV-19/Wales/MILK-9E2153/2020|EPI_ISL_581212|2020 ···················· ···················· ····················· ···················· ···················· ····················
L 4022 hCoV-19/England/MILK-9E1D32/2020|EPI_ISL_581213|20 ···················· ···················· ····················· ···················· ···················· ····················
L 4023 hCoV-19/England/QEUH-9DECC2/2020|EPI_ISL_581214|20 ···················· ···················· ····················· ···················· ······T············· ····················
L 4024 hCoV-19/England/MILK-9E1D9C/2020|EPI_ISL_581215|20 ···················· ···················· ····················· ···················· ···················· ····················
L 4025 hCoV-19/England/MILK-9E25EE/2020|EPI_ISL_581216|20 ···················· ···················· ····················· ···················· ···················· ····················
L 4026 hCoV-19/England/QEUH-9DEEDB/2020|EPI_ISL_581217|20 ···················· ···················· ····················· ···················· ···················· ····················
L 4027 hCoV-19/England/MILK-9E2997/2020|EPI_ISL_581218|20 ···················· ····A··············· ····················· ···················· ···················· ····················
L 4028 hCoV-19/England/MILK-9E2F0E/2020|EPI_ISL_581219|20 ···················· ···················· ····················· ···················· ···················· ····················
L 4029 hCoV-19/England/MILK-9E1D23/2020|EPI_ISL_581220|20 ···················· ···················· ····················· ···················· ···················· ····················
L 4030 hCoV-19/England/MILK-9E2F68/2020|EPI_ISL_581221|20 ···················· ···················· ····················· ···················· ···················· ····················
L 4031 hCoV-19/Wales/MILK-9E3213/2020|EPI_ISL_581222|2020 ···················· ···················· ····················· ···················· ···················· ····················
L 4032 hCoV-19/England/QEUH-9DE9BC/2020|EPI_ISL_581223|20 ···················· ···················· ····················· ···················· ···················· ····················
L 4033 hCoV-19/England/MILK-9E3161/2020|EPI_ISL_581224|20 ···················· ···················· ····················· ···················· ···················· ····················
L 4034 hCoV-19/Wales/MILK-9E227E/2020|EPI_ISL_581225|2020 ···················· ···················· ····················· ···················· ···················· ····················
L 4035 hCoV-19/England/QEUH-9DE785/2020|EPI_ISL_581226|20 ···················· ·A·················· ····················· ···················· ···················· ··A·················
L 4036 hCoV-19/England/MILK-9E2B55/2020|EPI_ISL_581227|20 ···················· ···················· ····················· ···················· ···················· ····················
L 4037 hCoV-19/England/MILK-9E1DAB/2020|EPI_ISL_581228|20 ···················· ···················· ····················· ···················· ···················· ····················
L 4038 hCoV-19/England/QEUH-9DE776/2020|EPI_ISL_581229|20 ···················· ···················· ····················· ···················· ···················· ····················
L 4039 hCoV-19/England/QEUH-9DE837/2020|EPI_ISL_581230|20 ···················· ···················· ····················· ···················· ···················· ····················
L 4040 hCoV-19/England/MILK-9E2C52/2020|EPI_ISL_581231|20 ···················· ···················· ····················· ···················· ···················· ····················
L 4041 hCoV-19/Wales/MILK-9E325F/2020|EPI_ISL_581232|2020 ···················· ···················· ····················· ···················· ···················· ····················
L 4042 hCoV-19/England/MILK-9E1DE7/2020|EPI_ISL_581233|20 ···················· ···················· ····················· ···················· ···················· ····················
L 4043 hCoV-19/Wales/MILK-9E2B64/2020|EPI_ISL_581234|2020 ···················· ·········A·········· ····················· ···················· ···················· ····················
L 4044 hCoV-19/England/MILK-9E344A/2020|EPI_ISL_581235|20 ···················· ···················· ····················· ···················· ···················· ····················
L 4045 hCoV-19/England/MILK-9E2FD1/2020|EPI_ISL_581236|20 ···················· ···················· ····················· ···················· ···················· ····················
L 4046 hCoV-19/England/MILK-9E2D8C/2020|EPI_ISL_581237|20 ···················· ···················· ····················· ···················· ···················· ····················
L 4047 hCoV-19/England/MILK-9E32E6/2020|EPI_ISL_581238|20 ···················· ···················· ····················· ···················· ···················· ····················
L 4048 hCoV-19/England/MILK-9E1D7E/2020|EPI_ISL_581239|20 ···················· ···················· ····················· ···················· ···················· ····················
L 4049 hCoV-19/England/MILK-9E1F4B/2020|EPI_ISL_581240|20 ···················· ···················· ····················· ···················· ···················· ····················
L 4050 hCoV-19/England/MILK-9E33A7/2020|EPI_ISL_581241|20 ···················· ·A·················· ····················· ···················· ···················· ··A·················
L 4051 hCoV-19/Wales/MILK-9E2038/2020|EPI_ISL_581243|2020 ···················· ···················· ····················· ···················· ···················· ····················
L 4052 hCoV-19/England/MILK-9E1F87/2020|EPI_ISL_581244|20 ···················· ···················· ····················· ···················· ···················· ····················
L 4053 hCoV-19/England/QEUH-9DE5E5/2020|EPI_ISL_581245|20 ···················· ···················· ····················· ···················· ···················· ····················
L 4054 hCoV-19/England/MILK-9E3064/2020|EPI_ISL_581246|20 ···················· ···················· ····················· ···················· ···················· ····················
L 4055 hCoV-19/England/QEUH-9DE5B8/2020|EPI_ISL_581247|20 ···················· ···················· ····················· ···················· ···················· ····················
L 4056 hCoV-19/England/MILK-9E2FC2/2020|EPI_ISL_581248|20 ···················· ···················· ····················· ···················· ···················· ····················
L 4057 hCoV-19/England/MILK-9E25FD/2020|EPI_ISL_581249|20 ···················· ···················· ····················· ···················· ···················· ····················
L 4058 hCoV-19/Wales/MILK-9E3204/2020|EPI_ISL_581250|2020 ···················· ···················· ····················· ···················· ···················· ····················
L 4059 hCoV-19/England/MILK-9E3091/2020|EPI_ISL_581251|20 ···················· ···················· ····················· ···················· ······T············· ····················
L 4060 hCoV-19/England/MILK-9E33C5/2020|EPI_ISL_581252|20 ···················· ···················· ····················· ···················· ···················· ····················
L 4061 hCoV-19/England/MILK-9AAE05/2020|EPI_ISL_581390|20 ···················· ···················· ····················· ···················· ······T············· ····················
L 4062 hCoV-19/England/QEUH-99817A/2020|EPI_ISL_581392|20 ···················· ···················· ····················· ···················· ···················· ····················
L 4063 hCoV-19/England/CAMC-991144/2020|EPI_ISL_581394|20 ···················· ···················· ····················· ···················· ···················· ····················
L 4064 hCoV-19/Canada/MB-NML-1186/2020|EPI_ISL_582500|202 ···················· ·A·················· ····················· ···················· ···················· ··A·················
L 4065 hCoV-19/Canada/MB-NML-1187/2020|EPI_ISL_582501|202 ···················· ·A·················· ····················· ···················· ···················· ··A·················
L 4066 hCoV-19/Canada/MB-NML-1188/2020|EPI_ISL_582502|202 ···················· ·A·················· ····················· ···················· ···················· ··A·················
L 4067 hCoV-19/Canada/MB-NML-1189/2020|EPI_ISL_582503|202 ···················· ···················· ····················· ···················· ···················· ····················
L 4068 hCoV-19/Canada/MB-NML-1190/2020|EPI_ISL_582504|202 ···················· ·A·················· ····················· ···················· ···················· ····················
L 4069 hCoV-19/Canada/MB-NML-1191/2020|EPI_ISL_582505|202 ···················· ···················· ····················· ···················· ···················· ····················

| | | | | | | | |
|---|---|---|---|---|---|---|---|
| L | 4070 hCoV-19/Canada/MB-NML-1193/2020\|EPI_ISL_582506\|202 | ···················· | ···················· | ···················· | ···················· | ···················· | ···················· |
| L | 4071 hCoV-19/Canada/MB-NML-1195/2020\|EPI_ISL_582507\|202 | ···················· | ···················· | ···················· | ···················· | ···················· | ···················· |
| L | 4072 hCoV-19/France/ARA-CDP29289/2020\|EPI_ISL_582508\|20 | ···················· | ····A··············· | ···················· | ···················· | ···················· | ···················· |
| L | 4073 hCoV-19/Ukraine/Kharkiv-817/2020\|EPI_ISL_582509\|20 | ···················· | ···················· | ···················· | ···················· | ···················· | ···················· |
| L | 4074 hCoV-19/Ukraine/Kharkiv-877/2020\|EPI_ISL_582510\|20 | ···················· | ···················· | ···················· | ···················· | ···················· | ···················· |
| L | 4075 hCoV-19/Ukraine/Kharkiv-879/2020\|EPI_ISL_582511\|20 | ···················· | ···················· | ···················· | ···················· | ···················· | ···················· |
| L | 4076 hCoV-19/Ukraine/Kharkiv-882/2020\|EPI_ISL_582512\|20 | ···················· | ···················· | ···················· | ···················· | ···················· | ···················· |
| L | 4077 hCoV-19/Ukraine/Kharkiv-782/2020\|EPI_ISL_582513\|20 | ···················· | ···················· | ···················· | ···················· | ···················· | ···················· |
| L | 4078 hCoV-19/Portugal/PT0939/2020\|EPI_ISL_582514\|2020-0 | ···················· | ···················· | ···················· | ···················· | ···················· | ···················· |
| L | 4079 hCoV-19/Portugal/PT0954/2020\|EPI_ISL_582515\|2020-0 | ···················· | ···················· | ···················· | ···················· | ···················· | ···················· |
| L | 4080 hCoV-19/Portugal/PT0982/2020\|EPI_ISL_582516\|2020-0 | ···················· | ···················· | ···················· | ···················· | ···················· | ···················· |
| L | 4081 hCoV-19/Portugal/PT1003/2020\|EPI_ISL_582517\|2020-0 | ···················· | ···················· | ···················· | ···················· | ···················· | ···················· |
| L | 4082 hCoV-19/Portugal/PT1164/2020\|EPI_ISL_582518\|2020-0 | ···················· | ···················· | ···················· | ···················· | ···················· | ···················· |
| L | 4083 hCoV-19/Portugal/PT1179/2020\|EPI_ISL_582519\|2020-0 | ···················· | ···················· | ···················· | ···················· | ···················· | ···················· |
| L | 4084 hCoV-19/Portugal/PT1295/2020\|EPI_ISL_582522\|2020-0 | ···················· | ···················· | ···················· | ···················· | ···················· | ···················· |
| L | 4085 hCoV-19/Portugal/PT1297/2020\|EPI_ISL_582523\|2020-0 | ···················· | ···················· | ···················· | ···················· | ···················· | ···················· |
| L | 4086 hCoV-19/Serbia/Kragujevac-00040304/2020\|EPI_ISL_58 | ···················· | ···················· | ···················· | ···················· | ···················· | ···················· |
| L | 4087 hCoV-19/Serbia/Cuprija-00880404/2020\|EPI_ISL_58252 | ···················· | ···················· | ···················· | ···················· | ···················· | ···················· |
| L | 4088 hCoV-19/Serbia/Krusevac-02260604/2020\|EPI_ISL_5825 | ···················· | ···················· | ···················· | ···················· | ···················· | ···················· |
| L | 4089 hCoV-19/Serbia/Krusevac-05881304/2020\|EPI_ISL_5825 | ···················· | ···················· | ···················· | ···················· | ···················· | ···················· |
| L | 4090 hCoV-19/Serbia/Krusevac-07961604/2020\|EPI_ISL_5825 | ···················· | ···················· | ···················· | ···················· | ···················· | ···················· |
| L | 4091 hCoV-19/Serbia/Krusevac-07711704/2020\|EPI_ISL_5825 | ···················· | ···················· | ···················· | ···················· | ···················· | ···················· |
| L | 4092 hCoV-19/Serbia/Kosovska Mitrovica-08332107/2020\|EP | ···················· | ···················· | ···················· | ···················· | ···················· | ···················· |
| L | 4093 hCoV-19/Serbia/Kraljevo-01242507/2020\|EPI_ISL_5825 | ···················· | ···················· | ···················· | ···················· | ···················· | ···················· |
| L | 4094 hCoV-19/Serbia/Kraljevo-01452507/2020\|EPI_ISL_5825 | ···················· | ···················· | ···················· | ···················· | ···················· | ···················· |
| L | 4095 hCoV-19/Serbia/Kraljevo-02092507/2020\|EPI_ISL_5825 | ···················· | ···················· | ···················· | ···················· | ···················· | ···················· |
| L | 4096 hCoV-19/Serbia/Kraljevo-03762507/2020\|EPI_ISL_5825 | ···················· | ···················· | ···················· | ···················· | ···················· | ···················· |
| L | 4097 hCoV-19/USA/TX-QDX-984/2020\|EPI_ISL_571800\|2020-03 | ···················· | ···················· | ···················· | ···················· | ···················· | ···················· |
| L | 4098 hCoV-19/USA/WA-QDX-1058/2020\|EPI_ISL_571801\|2020-0 | ···················· | ···················· | ···················· | ···················· | ···················· | ···················· |
| L | 4099 hCoV-19/USA/WA-QDX-1060/2020\|EPI_ISL_571802\|2020-0 | ···················· | ···················· | ···················· | ···················· | ···················· | ···················· |
| L | 4100 hCoV-19/USA/AZ-QDX-1079/2020\|EPI_ISL_571803\|2020-0 | ···················· | ···················· | ···················· | ···················· | ···················· | ···················· |
| L | 4101 hCoV-19/USA/CA-QDX-1064/2020\|EPI_ISL_571804\|2020-0 | ···················· | ···················· | ···················· | ···················· | ···················· | ···················· |
| L | 4102 hCoV-19/USA/CA-QDX-1065/2020\|EPI_ISL_571805\|2020-0 | ···················· | ···················· | ···················· | ···················· | ···················· | ···················· |
| L | 4103 hCoV-19/USA/CA-QDX-1067/2020\|EPI_ISL_571806\|2020-0 | ···················· | ···················· | ···················· | ···················· | ···················· | ···················· |
| L | 4104 hCoV-19/USA/CA-QDX-1068/2020\|EPI_ISL_571807\|2020-0 | ···················· | ···················· | ···················· | ···················· | ···················· | ···················· |
| L | 4105 hCoV-19/USA/CA-QDX-1069/2020\|EPI_ISL_571808\|2020-0 | ···················· | ···················· | ···················· | ···················· | ···················· | ···················· |
| L | 4106 hCoV-19/USA/CA-QDX-1095/2020\|EPI_ISL_571809\|2020-0 | ···················· | ···················· | ···················· | ···················· | ···················· | ···················· |
| L | 4107 hCoV-19/USA/CA-QDX-1102/2020\|EPI_ISL_571810\|2020-0 | ···················· | ···················· | ···················· | ···················· | ···················· | ···················· |
| L | 4108 hCoV-19/USA/CA-QDX-1115/2020\|EPI_ISL_571811\|2020-0 | ···················· | ···················· | ···················· | ···················· | ···················· | ···················· |
| L | 4109 hCoV-19/USA/CA-QDX-1116/2020\|EPI_ISL_571812\|2020-0 | ···················· | ···················· | ···················· | ···················· | ···················· | ···················· |
| L | 4110 hCoV-19/USA/CA-QDX-1117/2020\|EPI_ISL_571813\|2020-0 | ···················· | ···················· | ···················· | ···················· | ···················· | ···················· |
| L | 4111 hCoV-19/USA/CA-QDX-1119/2020\|EPI_ISL_571814\|2020-0 | ···················· | ···················· | ···················· | ···················· | ···················· | ···················· |
| L | 4112 hCoV-19/USA/CA-QDX-1168/2020\|EPI_ISL_571815\|2020-0 | ···················· | ···················· | ···················· | ···················· | ···················· | ···················· |
| L | 4113 hCoV-19/USA/CA-QDX-1169/2020\|EPI_ISL_571816\|2020-0 | ···················· | ···················· | ···················· | ···················· | ···················· | ···················· |
| L | 4114 hCoV-19/USA/CA-QDX-1170/2020\|EPI_ISL_571817\|2020-0 | ···················· | ···················· | ···················· | ···················· | ···················· | ···················· |
| L | 4115 hCoV-19/USA/CA-QDX-1171/2020\|EPI_ISL_571818\|2020-0 | ···················· | ···················· | ···················· | ···················· | ···················· | ···················· |
| L | 4116 hCoV-19/USA/CA-QDX-1172/2020\|EPI_ISL_571819\|2020-0 | ···················· | ···················· | ···················· | ···················· | ···················· | ···················· |
| L | 4117 hCoV-19/USA/CA-QDX-1173/2020\|EPI_ISL_571820\|2020-0 | ···················· | ···················· | ···················· | ···················· | ···················· | ···················· |
| L | 4118 hCoV-19/USA/PA-QDX-1061/2020\|EPI_ISL_571821\|2020-0 | ···················· | ···················· | ···················· | ···················· | ···················· | ···················· |
| L | 4119 hCoV-19/USA/FL-QDX-1114/2020\|EPI_ISL_571822\|2020-0 | ···················· | ···················· | ···················· | ···················· | ···················· | ···················· |
| L | 4120 hCoV-19/USA/GA-QDX-1073/2020\|EPI_ISL_571823\|2020-0 | ···················· | ···················· | ···················· | ···················· | ···················· | ···················· |
| L | 4121 hCoV-19/USA/ID-QDX-1081/2020\|EPI_ISL_571824\|2020-0 | ···················· | ···················· | ···················· | ···················· | ···················· | ···················· |
| L | 4122 hCoV-19/USA/ID-QDX-1082/2020\|EPI_ISL_571825\|2020-0 | ···················· | ···················· | ···················· | ···················· | ···················· | ···················· |
| L | 4123 hCoV-19/USA/ID-QDX-1083/2020\|EPI_ISL_571826\|2020-0 | ···················· | ···················· | ···················· | ···················· | ···················· | ···················· |
| L | 4124 hCoV-19/USA/ID-QDX-1086/2020\|EPI_ISL_571827\|2020-0 | ···················· | ···················· | ···················· | ···················· | ···················· | ···················· |
| L | 4125 hCoV-19/USA/ID-QDX-1087/2020\|EPI_ISL_571828\|2020-0 | ···················· | ···················· | ···················· | ···················· | ···················· | ···················· |
| L | 4126 hCoV-19/USA/ID-QDX-1088/2020\|EPI_ISL_571829\|2020-0 | ···················· | ···················· | ···················· | ···················· | ···················· | ···················· |
| L | 4127 hCoV-19/USA/ID-QDX-1089/2020\|EPI_ISL_571830\|2020-0 | ···················· | ···················· | ···················· | ···················· | ···················· | ···················· |
| L | 4128 hCoV-19/USA/ID-QDX-1090/2020\|EPI_ISL_571831\|2020-0 | ···················· | ···················· | ···················· | ···················· | ···················· | ···················· |
| L | 4129 hCoV-19/USA/ID-QDX-1091/2020\|EPI_ISL_571832\|2020-0 | ···················· | ···················· | ···················· | ···················· | ···················· | ···················· |
| L | 4130 hCoV-19/USA/ID-QDX-1092/2020\|EPI_ISL_571833\|2020-0 | ···················· | ···················· | ···················· | ···················· | ···················· | ···················· |
| L | 4131 hCoV-19/USA/ID-QDX-1093/2020\|EPI_ISL_571834\|2020-0 | ···················· | ···················· | ···················· | ···················· | ···················· | ···················· |
| L | 4132 hCoV-19/USA/ID-QDX-1094/2020\|EPI_ISL_571835\|2020-0 | ···················· | ···················· | ···················· | ···················· | ···················· | ···················· |
| L | 4133 hCoV-19/USA/ID-QDX-1105/2020\|EPI_ISL_571836\|2020-0 | ···················· | ···················· | ···················· | ···················· | ···················· | ···················· |
| L | 4134 hCoV-19/USA/ID-QDX-1106/2020\|EPI_ISL_571837\|2020-0 | ···················· | ···················· | ···················· | ···················· | ···················· | ···················· |
| L | 4135 hCoV-19/USA/ID-QDX-1109/2020\|EPI_ISL_571838\|2020-0 | ···················· | ···················· | ···················· | ···················· | ···················· | ···················· |
| L | 4136 hCoV-19/USA/ID-QDX-1110/2020\|EPI_ISL_571839\|2020-0 | ···················· | ···················· | ···················· | ···················· | ···················· | ···················· |
| L | 4137 hCoV-19/USA/ID-QDX-1111/2020\|EPI_ISL_571840\|2020-0 | ···················· | ···················· | ···················· | ···················· | ···················· | ···················· |
| L | 4138 hCoV-19/USA/ID-QDX-1112/2020\|EPI_ISL_571841\|2020-0 | ···················· | ···················· | ···················· | ···················· | ···················· | ···················· |
| L | 4139 hCoV-19/USA/IL-QDX-1098/2020\|EPI_ISL_571842\|2020-0 | ···················· | ···················· | ···················· | ···················· | ···················· | ···················· |
| L | 4140 hCoV-19/USA/IL-QDX-1100/2020\|EPI_ISL_571843\|2020-0 | ···················· | ···················· | ···················· | ···················· | ···················· | ···················· |
| L | 4141 hCoV-19/USA/IL-QDX-1101/2020\|EPI_ISL_571844\|2020-0 | ···················· | ···················· | ···················· | ···················· | ···················· | ···················· |
| L | 4142 hCoV-19/USA/IL-QDX-1103/2020\|EPI_ISL_571845\|2020-0 | ···················· | ···················· | ···················· | ···················· | ···················· | ···················· |
| L | 4143 hCoV-19/USA/IL-QDX-1108/2020\|EPI_ISL_571846\|2020-0 | ···················· | ···················· | ···················· | ···················· | ···················· | ···················· |
| L | 4144 hCoV-19/USA/IL-QDX-1113/2020\|EPI_ISL_571847\|2020-0 | ···················· | ···················· | ···················· | ···················· | ···················· | ···················· |
| L | 4145 hCoV-19/USA/LA-QDX-1070/2020\|EPI_ISL_571848\|2020-0 | ···················· | ···················· | ···················· | ···················· | ···················· | ···················· |
| L | 4146 hCoV-19/USA/MI-QDX-1099/2020\|EPI_ISL_571849\|2020-0 | ···················· | ···················· | ···················· | ···················· | ···················· | ···················· |
| L | 4147 hCoV-19/Wales/PHWC-16C3AE/2020\|EPI_ISL_574048\|2020 | ···················· | ···················· | ···················· | ···················· | ···················· | ···················· |
| L | 4148 hCoV-19/Wales/PHWC-16C3BD/2020\|EPI_ISL_574049\|2020 | ···················· | ···················· | ···················· | ···················· | ···················· | ···················· |
| L | 4151 hCoV-19/Wales/PHWC-16C441/2020\|EPI_ISL_574056\|2020 | ···················· | ···················· | ···················· | ···················· | ···················· | ···················· |

| | | | | | | | | | |
|---|---|---|---|---|---|---|---|---|---|
| L | 4152 | hCoV-19/Wales/PHWC-16C48D/2020\|EPI_ISL_574058\|2020 | ···················· | ···················· | ···················· | ···················· | ···················· | ···················· | |
| L | 4154 | hCoV-19/Wales/PHWC-16C5C6/2020\|EPI_ISL_574071\|2020 | ···················· | ···················· | ···················· | ···················· | ···················· | ···················· | |
| L | 4155 | hCoV-19/Wales/PHWC-16C5D5/2020\|EPI_ISL_574072\|2020 | ···················· | ···················· | ···················· | ···················· | ···················· | ···················· | |
| L | 4156 | hCoV-19/Wales/PHWC-16C5F3/2020\|EPI_ISL_574073\|2020 | ···················· | ···················· | ···················· | ···················· | ···················· | ···················· | |
| L | 4157 | hCoV-19/Wales/PHWC-16C60F/2020\|EPI_ISL_574074\|2020 | ···················· | ···················· | ···················· | ···················· | ···················· | ···················· | |
| L | 4158 | hCoV-19/Wales/PHWC-16C61E/2020\|EPI_ISL_574075\|2020 | ···················· | ···················· | ···················· | ···················· | ···················· | ···················· | |
| L | 4159 | hCoV-19/Wales/PHWC-16C62D/2020\|EPI_ISL_574076\|2020 | ···················· | ···················· | ···················· | ···················· | ···················· | ···················· | |
| L | 4160 | hCoV-19/Wales/PHWC-16C63C/2020\|EPI_ISL_574077\|2020 | ···················· | ···················· | ···················· | ···················· | ···················· | ···················· | |
| L | 4161 | hCoV-19/Wales/PHWC-16C64B/2020\|EPI_ISL_574078\|2020 | ···················· | ···················· | ···················· | ···················· | ···················· | ···················· | |
| L | 4163 | hCoV-19/Wales/PHWC-16C669/2020\|EPI_ISL_574080\|2020 | ···················· | ···················· | ···················· | ···················· | ···················· | ···················· | |
| L | 4164 | hCoV-19/Wales/PHWC-16C678/2020\|EPI_ISL_574081\|2020 | ···················· | ···················· | ···················· | ···················· | ···················· | ···················· | |
| L | 4165 | hCoV-19/Wales/PHWC-16C6A5/2020\|EPI_ISL_574082\|2020 | ···················· | ···················· | ···················· | ···················· | ···················· | ···················· | |
| L | 4166 | hCoV-19/Wales/PHWC-16C6B4/2020\|EPI_ISL_574083\|2020 | ···················· | ···················· | ···················· | ···················· | ···················· | ···················· | |
| L | 4167 | hCoV-19/Wales/PHWC-16C6C3/2020\|EPI_ISL_574084\|2020 | ···················· | ···················· | ···················· | ···················· | ···················· | ···················· | |
| L | 4168 | hCoV-19/Wales/PHWC-16C6F0/2020\|EPI_ISL_574085\|2020 | ···················· | ···················· | ···················· | ···················· | ···················· | ···················· | |
| L | 4169 | hCoV-19/Wales/PHWC-16C72A/2020\|EPI_ISL_574086\|2020 | ···················· | ···················· | ···················· | ···················· | ···················· | ···················· | |
| L | 4170 | hCoV-19/Wales/PHWC-16C739/2020\|EPI_ISL_574087\|2020 | ···················· | ···················· | ···················· | ···················· | ···················· | ···················· | |
| L | 4171 | hCoV-19/Wales/PHWC-16C748/2020\|EPI_ISL_574088\|2020 | ···················· | ···················· | ···················· | ···················· | ···················· | ···················· | |
| L | 4172 | hCoV-19/Wales/PHWC-16C775/2020\|EPI_ISL_574089\|2020 | ···················· | ···················· | ···················· | ···················· | ···················· | ···················· | |
| L | 4173 | hCoV-19/Wales/PHWC-16C784/2020\|EPI_ISL_574090\|2020 | ···················· | ···················· | ···················· | ···················· | ···················· | ···················· | |
| L | 4174 | hCoV-19/Wales/PHWC-16C793/2020\|EPI_ISL_574091\|2020 | ···················· | ···················· | ···················· | ···················· | ···················· | ···················· | |
| L | 4175 | hCoV-19/Wales/PHWC-16C7A2/2020\|EPI_ISL_574092\|2020 | ···················· | ···················· | ···················· | ···················· | ···················· | ···················· | |
| L | 4176 | hCoV-19/Wales/PHWC-16C7B1/2020\|EPI_ISL_574093\|2020 | ···················· | ···················· | ···················· | ···················· | ···················· | ···················· | |
| L | 4177 | hCoV-19/Wales/PHWC-16C7C0/2020\|EPI_ISL_574094\|2020 | ···················· | ···················· | ···················· | ···················· | ···················· | ···················· | |
| L | 4178 | hCoV-19/Wales/PHWC-16C7DF/2020\|EPI_ISL_574095\|2020 | ···················· | ···················· | ···················· | ···················· | ···················· | ···················· | |
| L | 4179 | hCoV-19/Wales/PHWC-16C7EE/2020\|EPI_ISL_574096\|2020 | ···················· | ···················· | ···················· | ···················· | NNN················· | ···················· | 1 |
| L | 4181 | hCoV-19/Wales/PHWC-16C809/2020\|EPI_ISL_574098\|2020 | ···················· | ···················· | ···················· | ···················· | ···················· | ···················· | |
| L | 4182 | hCoV-19/Wales/PHWC-16C818/2020\|EPI_ISL_574099\|2020 | ···················· | ···················· | ···················· | ···················· | ···················· | ···················· | |
| L | 4183 | hCoV-19/Wales/PHWC-16C836/2020\|EPI_ISL_574100\|2020 | ···················· | ···················· | ···················· | ···················· | ···················· | ···················· | |
| L | 4184 | hCoV-19/Wales/PHWC-16C845/2020\|EPI_ISL_574101\|2020 | ···················· | ···················· | ···················· | ···················· | ···················· | ···················· | |
| L | 4185 | hCoV-19/Wales/PHWC-16C854/2020\|EPI_ISL_574102\|2020 | ···················· | ···················· | ···················· | ···················· | ···················· | ···················· | |
| L | 4186 | hCoV-19/Wales/PHWC-16C872/2020\|EPI_ISL_574103\|2020 | ···················· | ···················· | ···················· | ···················· | ···················· | ···················· | |
| L | 4187 | hCoV-19/Wales/PHWC-16C881/2020\|EPI_ISL_574104\|2020 | ···················· | ···················· | ···················· | ···················· | ···················· | ···················· | |
| L | 4188 | hCoV-19/Wales/PHWC-16C890/2020\|EPI_ISL_574105\|2020 | ···················· | ···················· | ···················· | ···················· | ···················· | ···················· | |
| L | 4190 | hCoV-19/Wales/PHWC-16C8CD/2020\|EPI_ISL_574107\|2020 | ···················· | ···················· | ···················· | ···················· | ···················· | ···················· | |
| L | 4191 | hCoV-19/Wales/PHWC-16C8DC/2020\|EPI_ISL_574108\|2020 | ···················· | ···················· | ···················· | ···················· | ···················· | ···················· | |
| L | 4192 | hCoV-19/Wales/PHWC-16C8FA/2020\|EPI_ISL_574109\|2020 | ···················· | ···················· | ···················· | ···················· | ···················· | ···················· | |
| L | 4193 | hCoV-19/Wales/PHWC-16C915/2020\|EPI_ISL_574110\|2020 | ···················· | ···················· | ···················· | ···················· | ···················· | ···················· | |
| L | 4194 | hCoV-19/Wales/PHWC-16C924/2020\|EPI_ISL_574111\|2020 | ···················· | ···················· | ···················· | ···················· | ···················· | ···················· | |
| L | 4196 | hCoV-19/Wales/PHWC-16C942/2020\|EPI_ISL_574113\|2020 | ···················· | ···················· | ···················· | ···················· | ···················· | ···················· | |
| L | 4197 | hCoV-19/Wales/PHWC-16C960/2020\|EPI_ISL_574114\|2020 | ···················· | ···················· | ···················· | ···················· | ···················· | ···················· | |
| L | 4198 | hCoV-19/Wales/PHWC-16C9AC/2020\|EPI_ISL_574115\|2020 | ···················· | ···················· | ···················· | ···················· | ···················· | ···················· | |
| L | 4199 | hCoV-19/Wales/PHWC-16C9BB/2020\|EPI_ISL_574116\|2020 | ···················· | ···················· | ···················· | ···················· | ···················· | ···················· | |
| L | 4200 | hCoV-19/Wales/PHWC-16C9CA/2020\|EPI_ISL_574117\|2020 | ···················· | ···················· | ···················· | ···················· | ···················· | ···················· | |
| L | 4202 | hCoV-19/Wales/PHWC-16C9F7/2020\|EPI_ISL_574119\|2020 | ···················· | ···················· | ···················· | ···················· | ···················· | ···················· | |
| L | 4203 | hCoV-19/Wales/PHWC-16CA03/2020\|EPI_ISL_574120\|2020 | ···················· | ···················· | ···················· | ···················· | ···················· | ···················· | |
| L | 4204 | hCoV-19/Wales/PHWC-16CA12/2020\|EPI_ISL_574121\|2020 | ···················· | ···················· | ···················· | ···················· | ···················· | ···················· | |
| L | 4205 | hCoV-19/Wales/PHWC-16CA21/2020\|EPI_ISL_574122\|2020 | ···················· | ···················· | ···················· | ···················· | ···················· | ···················· | |
| L | 4206 | hCoV-19/Wales/PHWC-16CA30/2020\|EPI_ISL_574123\|2020 | ···················· | ···················· | ···················· | ···················· | ···················· | ···················· | |
| L | 4207 | hCoV-19/Wales/PHWC-16CA4F/2020\|EPI_ISL_574124\|2020 | ···················· | ···················· | ···················· | ···················· | ···················· | ···················· | |
| L | 4208 | hCoV-19/Wales/PHWC-16CA5E/2020\|EPI_ISL_574125\|2020 | ···················· | ···················· | ···················· | ···················· | ···················· | ···················· | |
| L | 4209 | hCoV-19/Wales/PHWC-16CA6D/2020\|EPI_ISL_574126\|2020 | ···················· | ···················· | ···················· | ···················· | ···················· | ···················· | |
| L | 4210 | hCoV-19/Wales/PHWC-16CA7C/2020\|EPI_ISL_574127\|2020 | ···················· | ···················· | ···················· | ···················· | ···················· | ···················· | |
| L | 4211 | hCoV-19/Wales/PHWC-16CA8B/2020\|EPI_ISL_574128\|2020 | ···················· | ···················· | ···················· | ···················· | ···················· | ···················· | |
| L | 4215 | hCoV-19/Wales/PHWC-16CAC7/2020\|EPI_ISL_574132\|2020 | ···················· | ···················· | ···················· | ···················· | ···················· | ···················· | |
| L | 4216 | hCoV-19/Wales/PHWC-16CAD6/2020\|EPI_ISL_574133\|2020 | ···················· | ···················· | ···················· | ···················· | ···················· | ···················· | |
| L | 4217 | hCoV-19/Wales/PHWC-16CAE5/2020\|EPI_ISL_574134\|2020 | ···················· | ···················· | ···················· | ···················· | ···················· | ···················· | |
| L | 4218 | hCoV-19/Wales/PHWC-16CAF4/2020\|EPI_ISL_574135\|2020 | ···················· | ···················· | ···················· | ···················· | ···················· | ···················· | |
| L | 4219 | hCoV-19/Wales/PHWC-16CB00/2020\|EPI_ISL_574136\|2020 | ···················· | ···················· | ···················· | ···················· | ···················· | ···················· | |
| L | 4220 | hCoV-19/Wales/PHWC-16CB4C/2020\|EPI_ISL_574137\|2020 | ···················· | ···················· | ···················· | ···················· | ···················· | ···················· | |
| L | 4221 | hCoV-19/Wales/PHWC-16CB5B/2020\|EPI_ISL_574138\|2020 | ···················· | ···················· | ···················· | ···················· | ···················· | ···················· | |
| L | 4222 | hCoV-19/Wales/PHWC-16CB6A/2020\|EPI_ISL_574139\|2020 | ···················· | ···················· | ···················· | ···················· | ···················· | ···················· | |
| L | 4223 | hCoV-19/Wales/PHWC-16CB79/2020\|EPI_ISL_574140\|2020 | ···················· | ···················· | ···················· | ···················· | ···················· | ···················· | |
| L | 4224 | hCoV-19/Wales/PHWC-16CB88/2020\|EPI_ISL_574141\|2020 | ···················· | ···················· | ···················· | ···················· | ···················· | ···················· | |
| L | 4225 | hCoV-19/Wales/PHWC-16CB97/2020\|EPI_ISL_574142\|2020 | ···················· | ···················· | ···················· | ···················· | ···················· | ···················· | |
| L | 4226 | hCoV-19/Wales/PHWC-16CBB5/2020\|EPI_ISL_574143\|2020 | ···················· | ···················· | ···················· | ···················· | ···················· | ···················· | |
| L | 4227 | hCoV-19/Wales/PHWC-16CBC4/2020\|EPI_ISL_574144\|2020 | ···················· | ···················· | ···················· | ···················· | ···················· | ···················· | |
| L | 4228 | hCoV-19/Wales/PHWC-16CBD3/2020\|EPI_ISL_574145\|2020 | ···················· | ···················· | ···················· | ···················· | ···················· | ···················· | |
| L | 4229 | hCoV-19/Wales/PHWC-16CBE2/2020\|EPI_ISL_574146\|2020 | ···················· | ···················· | ···················· | ···················· | ···················· | ···················· | |
| L | 4230 | hCoV-19/Wales/PHWC-16CD37/2020\|EPI_ISL_574161\|2020 | ···················· | ···················· | ···················· | ···················· | ···················· | ···················· | |
| L | 4231 | hCoV-19/Wales/PHWC-16CD55/2020\|EPI_ISL_574162\|2020 | ···················· | ···················· | ···················· | ···················· | ···················· | ···················· | |
| L | 4232 | hCoV-19/Wales/PHWC-16CD64/2020\|EPI_ISL_574163\|2020 | ···················· | ···················· | ···················· | ···················· | ···················· | ···················· | |
| L | 4233 | hCoV-19/Wales/PHWC-16CD82/2020\|EPI_ISL_574164\|2020 | ···················· | ···················· | ···················· | ···················· | ···················· | ···················· | |
| L | 4234 | hCoV-19/Scotland/QEUH-9DBF53/2020\|EPI_ISL_575379\|2 | ···················· | ···················· | ···················· | ···················· | ···················· | ···················· | |
| L | 4235 | hCoV-19/Scotland/QEUH-9DBEB0/2020\|EPI_ISL_575380\|2 | ···················· | ···················· | ···················· | ···················· | ···················· | ···················· | |
| L | 4236 | hCoV-19/Scotland/QEUH-9DA7C5/2020\|EPI_ISL_575381\|2 | ···················· | ···················· | ···················· | ···················· | ···················· | ···················· | |
| L | 4237 | hCoV-19/Scotland/QEUH-9DC20E/2020\|EPI_ISL_575386\|2 | ···················· | ···················· | ···················· | ···················· | ···················· | ···················· | |
| L | 4238 | hCoV-19/Scotland/QEUH-9DBE92/2020\|EPI_ISL_575387\|2 | ···················· | ···················· | ···················· | ···················· | ···················· | ···················· | |
| L | 4239 | hCoV-19/Scotland/QEUH-9DA9DE/2020\|EPI_ISL_575388\|2 | ···················· | ···················· | ···················· | ···················· | ···················· | ···················· | |
| L | 4240 | hCoV-19/England/ALDP-9CAD02/2020\|EPI_ISL_575412\|20 | ···················· | ···················· | ···················· | ···················· | ···················· | ···················· | |

L 4241 hCoV-19/England/ALDP-9CB2C6/2020|EPI_ISL_575413|20 ···················· ···················· ····················· ···················· ···················· ····················
L 4242 hCoV-19/England/ALDP-9CAE5A/2020|EPI_ISL_575414|20 ···················· ···················· ····················· ···················· ···················· ····················
L 4243 hCoV-19/England/ALDP-9CADA8/2020|EPI_ISL_575415|20 ···················· ···················· ····················· ···················· ···················· ····················
L 4244 hCoV-19/England/ALDP-9CBC40/2020|EPI_ISL_575416|20 ···················· ···················· ····················· ···················· ···················· ····················
L 4245 hCoV-19/England/ALDP-9CB2D5/2020|EPI_ISL_575417|20 ···················· ···················· ····················· ···················· ···················· ····················
L 4246 hCoV-19/England/ALDP-9CAF48/2020|EPI_ISL_575418|20 ···················· ···················· ····················· ···················· ···················· ····················
L 4247 hCoV-19/England/ALDP-9CAFA2/2020|EPI_ISL_575419|20 ···················· ···················· ····················· ···················· ···················· ····················
L 4248 hCoV-19/England/ALDP-9CB202/2020|EPI_ISL_575421|20 ···················· ·A·················· ····················· ···················· ···················· ····················
L 4249 hCoV-19/England/ALDP-9CB949/2020|EPI_ISL_575422|20 ···················· ···················· ····················· ···················· ···················· ····················
L 4250 hCoV-19/England/ALDP-9CB378/2020|EPI_ISL_575424|20 ···················· ···················· ····················· ···················· ···················· ····················
L 4251 hCoV-19/England/ALDP-9CBE4A/2020|EPI_ISL_575425|20 ···················· ···················· ····················· ···················· ···················· ····················
L 4252 hCoV-19/England/ALDP-9CB581/2020|EPI_ISL_575426|20 ···················· ···················· ····················· ···················· ···················· ····················
L 4253 hCoV-19/England/ALDP-9CBE68/2020|EPI_ISL_575427|20 ···················· ·A·················· ····················· ···················· ···················· ··A·················
L 4254 hCoV-19/England/ALDP-9CA9A4/2020|EPI_ISL_575428|20 ···················· ···················· ····················· ···················· ···················· ····················
L 4255 hCoV-19/England/ALDP-9CBFED/2020|EPI_ISL_575429|20 ···················· ···················· ····················· ···················· ···················· ····················
L 4256 hCoV-19/England/ALDP-9CB387/2020|EPI_ISL_575430|20 ···················· ···················· ····················· ···················· ···················· ····················
L 4257 hCoV-19/England/ALDP-9CB721/2020|EPI_ISL_575431|20 ···················· ···················· ····················· ···················· ···················· ····················
L 4258 hCoV-19/England/ALDP-9CB8A6/2020|EPI_ISL_575433|20 ···················· ···················· ····················· ···················· ···················· ····················
L 4259 hCoV-19/England/ALDP-9CBD2F/2020|EPI_ISL_575434|20 ···················· ···················· ····················· ···················· ···················· ····················
L 4260 hCoV-19/England/ALDP-9CBF65/2020|EPI_ISL_575435|20 ···················· ···················· ····················· ···················· ···················· ····················
L 4261 hCoV-19/England/ALDP-9CB493/2020|EPI_ISL_575436|20 ···················· ···················· ····················· ···················· ···················· ····················
L 4262 hCoV-19/England/ALDP-9CAC50/2020|EPI_ISL_575437|20 ···················· ···················· ····················· ···················· ···················· ····················
L 4263 hCoV-19/England/ALDP-9CAB35/2020|EPI_ISL_575438|20 ···················· ···················· ····················· ···················· ···················· ····················
L 4264 hCoV-19/England/ALDP-9CB9EF/2020|EPI_ISL_575439|20 ···················· ···················· ····················· ···················· ···T················ ····················
L 4265 hCoV-19/England/ALDP-9CADB7/2020|EPI_ISL_575441|20 ···················· ···················· ····················· ···················· ···················· ····················
L 4266 hCoV-19/England/ALDP-9CBFB0/2020|EPI_ISL_575442|20 ···················· ···················· ····················· ···················· ···················· ····················
L 4267 hCoV-19/England/ALDP-9CB34B/2020|EPI_ISL_575444|20 ···················· ···················· ····················· ···················· ···················· ····················
L 4268 hCoV-19/England/ALDP-9CB7D6/2020|EPI_ISL_575445|20 ···················· ·A·················· ····················· ···················· ···················· ··A·················
L 4269 hCoV-19/England/ALDP-9CB897/2020|EPI_ISL_575446|20 ···················· ·A·················· ····················· ···················· ···················· ····················
L 4270 hCoV-19/England/ALDP-9CB211/2020|EPI_ISL_575447|20 ···················· ···················· ····················· ···················· ···················· ····················
L 4271 hCoV-19/England/ALDP-9CB91C/2020|EPI_ISL_575448|20 ···················· ···················· ····················· ···················· ···················· ····················
L 4272 hCoV-19/England/ALDP-9CBD5C/2020|EPI_ISL_575449|20 ···················· ···················· ····················· ···················· ···················· ····················
L 4273 hCoV-19/England/ALDP-9CAD3F/2020|EPI_ISL_575450|20 ···················· ···················· ····················· ···················· ···················· ····················
L 4274 hCoV-19/England/ALDP-9CB27B/2020|EPI_ISL_575451|20 ···················· ···················· ····················· ···················· ···················· ····················
L 4275 hCoV-19/England/ALDP-9CCC9A/2020|EPI_ISL_575452|20 ···················· ···················· ····················· ···················· ···················· ····················
L 4276 hCoV-19/England/ALDP-9CBF1A/2020|EPI_ISL_575453|20 ···················· ···················· ····················· ···················· ···················· ····················
L 4277 hCoV-19/England/ALDP-9CB976/2020|EPI_ISL_575454|20 ···················· ···················· ····················· ···················· ···················· ····················
L 4278 hCoV-19/England/ALDP-9CBBE9/2020|EPI_ISL_575455|20 ···················· ···················· ····················· ···················· ···················· ····················
L 4279 hCoV-19/England/ALDP-9CB0CC/2020|EPI_ISL_575456|20 ···················· ···················· ····················· ···················· ···················· ····················
L 4280 hCoV-19/England/ALDP-9CB81F/2020|EPI_ISL_575457|20 ···················· ···················· ····················· ···················· ···T················ ····················
L 4281 hCoV-19/England/ALDP-9CB40C/2020|EPI_ISL_575458|20 ···················· ···················· ····················· ···················· ···················· ····················
L 4282 hCoV-19/England/ALDP-9CAC14/2020|EPI_ISL_575459|20 ···················· ···················· ····················· ···················· ···················· ····················
L 4283 hCoV-19/England/ALDP-9CB41B/2020|EPI_ISL_575461|20 ···················· ·A·················· ····················· ···················· ···················· ····················
L 4284 hCoV-19/England/ALDP-9CBA19/2020|EPI_ISL_575462|20 ···················· ···················· ····················· ···················· ···················· ····················
L 4285 hCoV-19/England/ALDP-9CB633/2020|EPI_ISL_575463|20 ···················· ···················· ····················· ···················· ···················· ····················
L 4286 hCoV-19/England/ALDP-9CB141/2020|EPI_ISL_575464|20 ···················· ···················· ····················· ···················· ···················· ····················
L 4287 hCoV-19/England/ALDP-9CB92B/2020|EPI_ISL_575465|20 ···················· ···················· ····················· ···················· ···················· ····················
L 4288 hCoV-19/England/ALDP-9CA9B3/2020|EPI_ISL_575466|20 ···················· ···················· ····················· ···················· ···················· ····················
L 4289 hCoV-19/England/ALDP-9CBE1D/2020|EPI_ISL_575467|20 ···················· ···················· ····················· ···················· ···················· ····················
L 4290 hCoV-19/England/ALDP-9CABEA/2020|EPI_ISL_575468|20 ···················· ···················· ····················· ···················· ···················· ····················
L 4291 hCoV-19/England/ALDP-9CAACF/2020|EPI_ISL_575469|20 ···················· ···················· ····················· ···················· ···················· ····················
L 4292 hCoV-19/England/ALDP-9CAE3C/2020|EPI_ISL_575470|20 ···················· ···················· ····················· ···················· ···················· ····················
L 4293 hCoV-19/England/ALDP-9CB42A/2020|EPI_ISL_575471|20 ···················· ···················· ····················· ···················· ···················· ····················
L 4294 hCoV-19/England/ALDP-9CB888/2020|EPI_ISL_575472|20 ···················· ···················· ····················· ···················· ···················· ····················
L 4295 hCoV-19/England/ALDP-9CB28A/2020|EPI_ISL_575473|20 ···················· ···················· ····················· ···················· ···················· ····················
L 4296 hCoV-19/England/ALDP-9CB67F/2020|EPI_ISL_575474|20 ···················· ···················· ····················· ···················· ···················· ····················
L 4297 hCoV-19/England/ALDP-9CB4EE/2020|EPI_ISL_575476|20 ···················· ···················· ····················· ···················· ···················· ····················
L 4298 hCoV-19/England/ALDP-9CCCA9/2020|EPI_ISL_575477|20 ···················· ···················· ····················· ···················· ···················· ····················
L 4299 hCoV-19/England/ALDP-9CAE78/2020|EPI_ISL_575494|20 ···················· ···················· ····················· ···················· ···················· ····················
L 4300 hCoV-19/England/ALDP-9CBD10/2020|EPI_ISL_575495|20 ···················· ···················· ····················· ···················· ···················· ····················
L 4301 hCoV-19/England/ALDP-9D95BD/2020|EPI_ISL_576743|20 ···················· ···················· ····················· ···················· ···················· ····················
L 4302 hCoV-19/England/ALDP-9D95CC/2020|EPI_ISL_576744|20 ···················· ···················· ····················· ···················· ···················· ····················
L 4303 hCoV-19/England/ALDP-9D95DB/2020|EPI_ISL_576745|20 ···················· ···················· ····················· ···················· ···················· ····················
L 4304 hCoV-19/England/ALDP-9D95F9/2020|EPI_ISL_576746|20 ···················· ···················· ····················· ···················· ···················· ····················
L 4305 hCoV-19/England/ALDP-9D9605/2020|EPI_ISL_576747|20 ···················· ···················· ····················· ···················· ···················· ····················
L 4306 hCoV-19/England/ALDP-9D9614/2020|EPI_ISL_576748|20 ···················· ·A·················· ····················· ···················· ···················· ··A·················
L 4307 hCoV-19/England/ALDP-9D9632/2020|EPI_ISL_576750|20 ···················· ···················· ····················· ···················· ···················· ····················
L 4308 hCoV-19/England/ALDP-9D9641/2020|EPI_ISL_576751|20 ···················· ·A·················· ····················· ···················· ···················· ··A·················
L 4309 hCoV-19/England/ALDP-9D966F/2020|EPI_ISL_576752|20 ···················· ···················· ····················· ···················· ···················· ····················
L 4310 hCoV-19/England/ALDP-9D96D8/2020|EPI_ISL_576754|20 ···················· ···················· ····················· ···················· ···················· ····················
L 4311 hCoV-19/England/ALDP-9D96F6/2020|EPI_ISL_576755|20 ···················· ···················· ····················· ···················· ···················· ····················
L 4312 hCoV-19/England/ALDP-9D976C/2020|EPI_ISL_576757|20 ···················· ···················· ····················· ···················· ···················· ····················
L 4313 hCoV-19/England/ALDP-9D9799/2020|EPI_ISL_576758|20 ···················· ···················· ····················· ···················· ···················· ····················
L 4314 hCoV-19/England/ALDP-9D97A8/2020|EPI_ISL_576759|20 ···················· ···················· ····················· ···················· ···················· ····················
L 4315 hCoV-19/England/ALDP-9D97B7/2020|EPI_ISL_576760|20 ···················· ···················· ····················· ···················· ···················· ····················
L 4316 hCoV-19/England/ALDP-9D97D5/2020|EPI_ISL_576761|20 ···················· ···················· ····················· ···················· ···················· ····················
L 4317 hCoV-19/England/ALDP-9D97F3/2020|EPI_ISL_576762|20 ···················· ···················· ····················· ···················· ···················· ····················
L 4318 hCoV-19/England/ALDP-9D9869/2020|EPI_ISL_576763|20 ···················· ·A·················· ····················· ···················· ···················· ··A·················
L 4319 hCoV-19/England/ALDP-9D9887/2020|EPI_ISL_576764|20 ···················· ···················· ····················· ···················· ···················· ····················
L 4320 hCoV-19/England/ALDP-9D9896/2020|EPI_ISL_576765|20 ···················· ···················· ····················· ···················· ···················· ····················

| | | | | | | | | | |
|---|---|---|---|---|---|---|---|---|---|
| L | 4321 | hCoV-19/England/ALDP-9D98A5/2020\|EPI_ISL_576766\|20 | ···················· | ···················· | ···················· | ···················· | ···················· | ···················· | |
| L | 4322 | hCoV-19/England/ALDP-9D98D2/2020\|EPI_ISL_576768\|20 | ···················· | ···················· | ···················· | ···················· | ···················· | ···················· | |
| L | 4323 | hCoV-19/England/ALDP-9D98E1/2020\|EPI_ISL_576769\|20 | ···················· | ···················· | ···················· | ···················· | ···················· | ···················· | |
| L | 4324 | hCoV-19/England/ALDP-9D98F0/2020\|EPI_ISL_576770\|20 | ···················· | ···················· | ···················· | ···················· | ···················· | ···················· | |
| L | 4325 | hCoV-19/England/ALDP-9D990C/2020\|EPI_ISL_576771\|20 | ···················· | ···················· | ···················· | ···················· | ···················· | ···················· | |
| L | 4326 | hCoV-19/England/ALDP-9D991B/2020\|EPI_ISL_576772\|20 | ···················· | ···················· | ···················· | ···················· | ···················· | ···················· | |
| L | 4327 | hCoV-19/England/ALDP-9D992A/2020\|EPI_ISL_576773\|20 | ···················· | ···················· | ···················· | ···················· | ···················· | ···················· | |
| L | 4328 | hCoV-19/England/ALDP-9D9939/2020\|EPI_ISL_576774\|20 | ···················· | ···················· | ···················· | ···················· | ···················· | ···················· | |
| L | 4329 | hCoV-19/England/ALDP-9D9948/2020\|EPI_ISL_576775\|20 | ···················· | ···················· | ···················· | ···················· | ···················· | ···················· | |
| L | 4330 | hCoV-19/England/ALDP-9D9957/2020\|EPI_ISL_576776\|20 | ···················· | ···················· | ···················· | ···················· | ···················· | ···················· | |
| L | 4331 | hCoV-19/England/ALDP-9D9966/2020\|EPI_ISL_576777\|20 | ···················· | ···················· | ···················· | ···················· | ···················· | ···················· | |
| L | 4332 | hCoV-19/England/ALDP-9D9975/2020\|EPI_ISL_576778\|20 | ···················· | ···················· | ···················· | ···················· | ···················· | ···················· | |
| L | 4333 | hCoV-19/England/ALDP-9D9984/2020\|EPI_ISL_576779\|20 | ···················· | ···················· | ···················· | ···················· | ···················· | ···················· | |
| L | 4334 | hCoV-19/England/ALDP-9D9993/2020\|EPI_ISL_576780\|20 | ···················· | ···················· | ···················· | ···················· | ···················· | ···················· | |
| L | 4335 | hCoV-19/England/ALDP-9D99B1/2020\|EPI_ISL_576781\|20 | ···················· | ···················· | ···················· | ···················· | ···················· | ···················· | |
| L | 4336 | hCoV-19/England/ALDP-9D99DF/2020\|EPI_ISL_576782\|20 | ···················· | ···················· | ···················· | ···················· | ···················· | ···················· | |
| L | 4337 | hCoV-19/England/ALDP-9D99EE/2020\|EPI_ISL_576783\|20 | ···················· | ···················· | ···················· | ···················· | ···················· | ···················· | |
| L | 4338 | hCoV-19/England/ALDP-9D99FD/2020\|EPI_ISL_576784\|20 | ···················· | ···················· | ···················· | ···················· | ···················· | ···················· | |
| L | 4339 | hCoV-19/England/ALDP-9D9A09/2020\|EPI_ISL_576785\|20 | ···················· | ···················· | ···················· | ···················· | ···················· | ···················· | |
| L | 4340 | hCoV-19/England/ALDP-9D9A18/2020\|EPI_ISL_576786\|20 | ···················· | ···················· | ···················· | ···················· | ···················· | ···················· | |
| L | 4341 | hCoV-19/England/ALDP-9D9A63/2020\|EPI_ISL_576787\|20 | ···················· | ···················· | ···················· | ···················· | ···················· | ···················· | |
| L | 4342 | hCoV-19/England/ALDP-9D9A72/2020\|EPI_ISL_576788\|20 | ···················· | ···················· | ···················· | ···················· | ···················· | ···················· | |
| L | 4343 | hCoV-19/England/ALDP-9D9A81/2020\|EPI_ISL_576789\|20 | ···················· | ···················· | ···················· | ···················· | ···················· | ···················· | |
| L | 4344 | hCoV-19/England/ALDP-9D9A90/2020\|EPI_ISL_576790\|20 | ···················· | ···················· | ···················· | ···················· | ···················· | ···················· | |
| L | 4345 | hCoV-19/England/ALDP-9D9AFA/2020\|EPI_ISL_576792\|20 | ···················· | ···················· | ···················· | ···················· | ···················· | ···················· | |
| L | 4346 | hCoV-19/England/ALDP-9D9B06/2020\|EPI_ISL_576793\|20 | ···················· | ···················· | ···················· | ···················· | ···················· | ···················· | |
| L | 4347 | hCoV-19/England/ALDP-9D9B15/2020\|EPI_ISL_576794\|20 | ···················· | ···················· | ···················· | ···················· | ···················· | ···················· | |
| L | 4348 | hCoV-19/England/ALDP-9D9B51/2020\|EPI_ISL_576795\|20 | ···················· | ···················· | ···················· | ···················· | ···················· | ···················· | |
| L | 4349 | hCoV-19/England/ALDP-9D9B7F/2020\|EPI_ISL_576796\|20 | ···················· | ···················· | ···················· | ···················· | ···················· | ···················· | |
| L | 4350 | hCoV-19/England/ALDP-9D9B8E/2020\|EPI_ISL_576797\|20 | ···················· | ···················· | ···················· | ···················· | ···················· | ···················· | |
| L | 4351 | hCoV-19/England/ALDP-9D9B9D/2020\|EPI_ISL_576798\|20 | ···················· | ···················· | ···················· | ···················· | ···················· | ···················· | |
| L | 4352 | hCoV-19/England/ALDP-9D9BBB/2020\|EPI_ISL_576799\|20 | ···················· | ···················· | ···················· | ···················· | ···················· | ···················· | |
| L | 4353 | hCoV-19/England/QEUH-9AED6E/2020\|EPI_ISL_573500\|20 | ···················· | ···················· | ···················· | ···················· | ···················· | ···················· | |
| L | 4354 | hCoV-19/England/QEUH-9AED9B/2020\|EPI_ISL_573502\|20 | ···················· | ···················· | ···················· | ···················· | ···················· | ···················· | |
| L | 4355 | hCoV-19/England/QEUH-9AEDAA/2020\|EPI_ISL_573503\|20 | ···················· | ···················· | ···················· | ···················· | ···················· | ···················· | |
| L | 4356 | hCoV-19/England/QEUH-9AEDB9/2020\|EPI_ISL_573504\|20 | ···················· | ···················· | ···················· | ···················· | ···················· | ···················· | |
| L | 4357 | hCoV-19/England/QEUH-9AEDC8/2020\|EPI_ISL_573505\|20 | ···················· | ···················· | ···················· | ···················· | ···················· | ···················· | |
| L | 4358 | hCoV-19/England/QEUH-9AEDD7/2020\|EPI_ISL_573506\|20 | ···················· | ···················· | ···················· | ···················· | ···················· | ···················· | |
| L | 4359 | hCoV-19/England/QEUH-9AEE01/2020\|EPI_ISL_573507\|20 | ···················· | ···················· | ···················· | ···················· | ···················· | ···················· | |
| L | 4360 | hCoV-19/England/QEUH-9AEE10/2020\|EPI_ISL_573508\|20 | ···················· | ···················· | ···················· | ···················· | ···················· | ···················· | |
| L | 4361 | hCoV-19/England/QEUH-9AEE2F/2020\|EPI_ISL_573509\|20 | ···················· | ···················· | ···················· | ···················· | ···················· | ···················· | |
| L | 4363 | hCoV-19/Wales/PHWC-16CC0D/2020\|EPI_ISL_574148\|2020 | ···················· | ···················· | ···················· | ···················· | ···················· | ···················· | |
| L | 4364 | hCoV-19/Wales/PHWC-16CC1C/2020\|EPI_ISL_574149\|2020 | ···················· | ···················· | ···················· | ···················· | ···················· | ···················· | |
| L | 4365 | hCoV-19/Wales/PHWC-16CC3A/2020\|EPI_ISL_574150\|2020 | ···················· | ···················· | ···················· | ···················· | NNN················· | ···················· | 1 |
| L | 4366 | hCoV-19/Wales/PHWC-16CC49/2020\|EPI_ISL_574151\|2020 | ···················· | ···················· | ···················· | ···················· | ···················· | ···················· | |
| L | 4367 | hCoV-19/Wales/PHWC-16CC67/2020\|EPI_ISL_574152\|2020 | ···················· | ···················· | ···················· | ···················· | ···················· | ···················· | |
| L | 4368 | hCoV-19/Wales/PHWC-16CC76/2020\|EPI_ISL_574153\|2020 | ···················· | ···················· | ···················· | ···················· | ···················· | ···················· | |
| L | 4372 | hCoV-19/Wales/PHWC-16CCD0/2020\|EPI_ISL_574157\|2020 | ···················· | ···················· | ···················· | ···················· | ···················· | ···················· | |
| L | 4373 | hCoV-19/Wales/PHWC-16CD0A/2020\|EPI_ISL_574158\|2020 | ···················· | ···················· | ···················· | ···················· | ···················· | ···················· | |
| L | 4374 | hCoV-19/Wales/PHWC-16CD19/2020\|EPI_ISL_574159\|2020 | ···················· | ···················· | ···················· | ···················· | ···················· | ···················· | |
| L | 4376 | hCoV-19/Wales/PHWC-16CD91/2020\|EPI_ISL_574165\|2020 | ···················· | ···················· | ···················· | ···················· | ···················· | ···················· | |
| L | 4378 | hCoV-19/Wales/PHWC-16CDBF/2020\|EPI_ISL_574167\|2020 | ···················· | ···················· | ···················· | ···················· | ···················· | ···················· | |
| L | 4379 | hCoV-19/Wales/PHWC-16CDCE/2020\|EPI_ISL_574168\|2020 | ···················· | ···················· | ···················· | ···················· | ···················· | ···················· | |
| L | 4380 | hCoV-19/Switzerland/BE-UNIBE-160-0067/2020\|EPI_ISL | ···················· | ···················· | ···················· | ···················· | ···················· | ···················· | |
| L | 4381 | hCoV-19/Switzerland/BE-UNIBE-161-0117/2020\|EPI_ISL | ···················· | ···················· | ···················· | ···················· | ···················· | ···················· | |
| L | 4382 | hCoV-19/Switzerland/BE-UNIBE-162-0355/2020\|EPI_ISL | ···················· | ···················· | ···················· | ···················· | ···················· | ···················· | |
| L | 4383 | hCoV-19/Switzerland/BE-UNIBE-163-0431/2020\|EPI_ISL | ···················· | ···········A········ | ···················· | ···················· | ···················· | ···················· | |
| L | 4384 | hCoV-19/Switzerland/BE-UNIBE-164-0436/2020\|EPI_ISL | ···················· | ···················· | ···················· | ···················· | ···················· | ···················· | |
| L | 4385 | hCoV-19/Switzerland/BE-UNIBE-165-0657/2020\|EPI_ISL | ···················· | ···················· | ···················· | ···················· | ···················· | ···················· | |
| L | 4386 | hCoV-19/Switzerland/BE-UNIBE-166-0852/2020\|EPI_ISL | ···················· | ···················· | ···················· | ···················· | ···················· | ···················· | |
| L | 4387 | hCoV-19/Switzerland/BE-UNIBE-167-0863/2020\|EPI_ISL | ···················· | ···················· | ···················· | ···················· | ···················· | ···················· | |
| L | 4388 | hCoV-19/Switzerland/BE-UNIBE-168-1178/2020\|EPI_ISL | ···················· | ···················· | ···················· | ···················· | ···················· | ···················· | |
| L | 4389 | hCoV-19/Switzerland/BE-UNIBE-169-1508/2020\|EPI_ISL | ···················· | ···················· | ···················· | ···················· | ···················· | ···················· | |
| L | 4390 | hCoV-19/Switzerland/BE-UNIBE-170-1618/2020\|EPI_ISL | ···················· | ···················· | ···················· | ···················· | ···················· | ···················· | |
| L | 4391 | hCoV-19/Switzerland/BE-UNIBE-171-1643/2020\|EPI_ISL | ···················· | ···················· | ···················· | ···················· | ···················· | ···················· | |
| L | 4392 | hCoV-19/Switzerland/BE-UNIBE-172-1644/2020\|EPI_ISL | ···················· | ···················· | ···················· | ···················· | ···················· | ···················· | |
| L | 4393 | hCoV-19/Switzerland/BE-UNIBE-173-2392/2020\|EPI_ISL | ···················· | ·A·················· | ···················· | ···················· | ···················· | ···················· | |
| L | 4394 | hCoV-19/Switzerland/BE-UNIBE-174-2395/2020\|EPI_ISL | ···················· | ···················· | ···················· | ···················· | ···················· | ···················· | |
| L | 4395 | hCoV-19/Switzerland/BE-UNIBE-175-2461/2020\|EPI_ISL | ···················· | ···················· | ···················· | ···················· | ···················· | ···················· | |
| L | 4396 | hCoV-19/Switzerland/BE-UNIBE-176-2465/2020\|EPI_ISL | ···················· | ···················· | ···················· | ···················· | ···················· | ···················· | |
| L | 4397 | hCoV-19/Switzerland/BE-UNIBE-178-2905/2020\|EPI_ISL | ···················· | ···················· | ···················· | ···················· | ···················· | ···················· | |
| L | 4398 | hCoV-19/Switzerland/BE-UNIBE-179-2921/2020\|EPI_ISL | ···················· | ···················· | ···················· | ···················· | ···················· | ···················· | |
| L | 4399 | hCoV-19/Switzerland/BE-UNIBE-180-2946/2020\|EPI_ISL | ···················· | ···················· | ···················· | ···················· | ···················· | ···················· | |
| L | 4400 | hCoV-19/Switzerland/BE-UNIBE-181-3245/2020\|EPI_ISL | ···················· | ···················· | ···················· | ···················· | ···················· | ···················· | |
| L | 4401 | hCoV-19/Switzerland/BE-UNIBE-182-3534/2020\|EPI_ISL | ···················· | ···················· | ···················· | ···················· | ···················· | ···················· | |
| L | 4402 | hCoV-19/Switzerland/BE-UNIBE-183-3586/2020\|EPI_ISL | ···················· | ···················· | ···················· | ···················· | ···················· | ···················· | |
| L | 4403 | hCoV-19/Switzerland/BE-UNIBE-184-3592/2020\|EPI_ISL | ···················· | ···················· | ···················· | ···················· | ···················· | ···················· | |
| L | 4404 | hCoV-19/Switzerland/BE-UNIBE-185-3816/2020\|EPI_ISL | ···················· | ···················· | ···················· | ···················· | ···················· | ···················· | |
| L | 4405 | hCoV-19/Switzerland/BE-UNIBE-186-3956/2020\|EPI_ISL | ···················· | ···················· | ···················· | ···················· | ···················· | ···················· | |
| L | 4406 | hCoV-19/Switzerland/BE-UNIBE-187-3981/2020\|EPI_ISL | ···················· | ···················· | ···················· | ···················· | ···················· | ···················· | |

| | | | | | | | |
|---|---|---|---|---|---|---|---|
| L | 4407 hCoV-19/Switzerland/BE-UNIBE-188-3982/2020\|EPI_ISL | ···················· | ···················· | ···················· | ···················· | ···················· | ···················· |
| L | 4408 hCoV-19/Switzerland/BE-UNIBE-189-4085/2020\|EPI_ISL | ···················· | ···················· | ···················· | ···················· | ···················· | ···················· |
| L | 4409 hCoV-19/Switzerland/BE-UNIBE-190-4089/2020\|EPI_ISL | ···················· | ·A·················· | ···················· | ···················· | ···················· | ···················· |
| L | 4410 hCoV-19/Switzerland/BE-UNIBE-191-4171/2020\|EPI_ISL | ···················· | ···················· | ···················· | ···················· | ···················· | ···················· |
| L | 4411 hCoV-19/Switzerland/BE-UNIBE-192-4208/2020\|EPI_ISL | ···················· | ···················· | ···················· | ···················· | ···················· | ···················· |
| L | 4412 hCoV-19/Switzerland/BE-UNIBE-193-4523/2020\|EPI_ISL | ···················· | ···················· | ···················· | ···················· | ···················· | ···················· |
| L | 4413 hCoV-19/Switzerland/BE-UNIBE-194-4693/2020\|EPI_ISL | ···················· | ···················· | ···················· | ···················· | ···················· | ···················· |
| L | 4414 hCoV-19/Switzerland/BE-UNIBE-195-4728/2020\|EPI_ISL | ···················· | ···················· | ···················· | ···················· | ···················· | ···················· |
| L | 4415 hCoV-19/Switzerland/BE-UNIBE-197-5085/2020\|EPI_ISL | ···················· | ···················· | ···················· | ···················· | ···················· | ···················· |
| L | 4416 hCoV-19/Switzerland/BE-UNIBE-198-5301/2020\|EPI_ISL | ···················· | ···················· | ···················· | ···················· | ···················· | ···················· |
| L | 4417 hCoV-19/Switzerland/BE-UNIBE-199-5783/2020\|EPI_ISL | ···················· | ·A·················· | ···················· | ···················· | ···················· | ···················· |
| L | 4418 hCoV-19/Switzerland/BE-UNIBE-200-5795/2020\|EPI_ISL | ···················· | ···················· | ···················· | ···················· | ···················· | ···················· |
| L | 4419 hCoV-19/Switzerland/BE-UNIBE-201-6020/2020\|EPI_ISL | ···················· | ···················· | ···················· | ···················· | ···················· | ···················· |
| L | 4420 hCoV-19/England/ALDP-9CAB9F/2020\|EPI_ISL_575400\|20 | ···················· | ···················· | ···················· | ···················· | ···················· | ···················· |
| L | 4421 hCoV-19/England/ALDP-9CBDC5/2020\|EPI_ISL_575401\|20 | ···················· | ···················· | ···················· | ···················· | ···················· | ···················· |
| L | 4422 hCoV-19/England/ALDP-9CB080/2020\|EPI_ISL_575402\|20 | ···················· | ···················· | ···················· | ···················· | ···················· | ···················· |
| L | 4423 hCoV-19/England/ALDP-9CC043/2020\|EPI_ISL_575403\|20 | ···················· | ···················· | ···················· | ···················· | ···················· | ···················· |
| L | 4424 hCoV-19/England/ALDP-9CBB8F/2020\|EPI_ISL_575406\|20 | ···················· | ···················· | ···················· | ···················· | ···················· | ···················· |
| L | 4425 hCoV-19/England/ALDP-9CBE77/2020\|EPI_ISL_575407\|20 | ···················· | ···················· | ···················· | ···················· | ···················· | ···················· |
| L | 4426 hCoV-19/England/ALDP-9CAD7B/2020\|EPI_ISL_575408\|20 | ···················· | ·A·················· | ···················· | ···················· | ···················· | ···A················ |
| L | 4427 hCoV-19/England/ALDP-9CB82E/2020\|EPI_ISL_575409\|20 | ···················· | ···················· | ···················· | ···················· | ···················· | ···················· |
| L | 4428 hCoV-19/England/ALDP-9CBC6E/2020\|EPI_ISL_575410\|20 | ···················· | ·A·······A·········· | ···················· | ···················· | ···················· | ···················· |
| L | 4429 hCoV-19/England/ALDP-9CABDB/2020\|EPI_ISL_575411\|20 | ···················· | ···················· | ···················· | ···················· | ···················· | ···················· |
| L | 4430 hCoV-19/England/ALDP-9CADE4/2020\|EPI_ISL_575478\|20 | ···················· | ···················· | ···················· | ···················· | ···················· | ···················· |
| L | 4431 hCoV-19/England/ALDP-9CAB80/2020\|EPI_ISL_575479\|20 | ···················· | ···················· | ···················· | ···················· | ···················· | ···················· |
| L | 4432 hCoV-19/England/ALDP-9CADC6/2020\|EPI_ISL_575480\|20 | ···················· | ···················· | ···················· | ···················· | ···················· | ···················· |
| L | 4433 hCoV-19/England/ALDP-9CB4B1/2020\|EPI_ISL_575481\|20 | ···················· | ···················· | ···················· | ···················· | ···················· | ···················· |
| L | 4434 hCoV-19/England/ALDP-9CAE0F/2020\|EPI_ISL_575482\|20 | ···················· | ···················· | ···················· | ···················· | ···················· | ···················· |
| L | 4435 hCoV-19/England/ALDP-9CB536/2020\|EPI_ISL_575483\|20 | ···················· | ······A············· | ···················· | ···················· | ···················· | ···················· |
| L | 4436 hCoV-19/England/ALDP-9CBF0B/2020\|EPI_ISL_575484\|20 | ···················· | ···················· | ···················· | ···················· | ···················· | ···················· |
| L | 4437 hCoV-19/England/ALDP-9CBDF2/2020\|EPI_ISL_575485\|20 | ···················· | ···················· | ···················· | ···················· | ···················· | ···················· |
| L | 4438 hCoV-19/England/ALDP-9CBBDA/2020\|EPI_ISL_575488\|20 | ···················· | ···················· | ···················· | ···················· | ···················· | ···················· |
| L | 4439 hCoV-19/England/ALDP-9CA9E0/2020\|EPI_ISL_575489\|20 | ···················· | ···················· | ···················· | ···················· | ···················· | ···················· |
| L | 4440 hCoV-19/England/ALDP-9CB9C1/2020\|EPI_ISL_575490\|20 | ···················· | ···················· | ···················· | ···················· | ···················· | ···················· |
| L | 4441 hCoV-19/England/ALDP-9CC09E/2020\|EPI_ISL_575491\|20 | ···················· | ···················· | ···················· | ···················· | ···················· | ···················· |
| L | 4442 hCoV-19/Wales/ALDP-9CB2F3/2020\|EPI_ISL_575496\|2020 | ···················· | ···················· | ···················· | ···················· | ···················· | ···················· |
| L | 4443 hCoV-19/England/ALDP-9CB9FE/2020\|EPI_ISL_575497\|20 | ···················· | ···················· | ···················· | ···················· | ···················· | ···················· |
| L | 4444 hCoV-19/England/ALDP-9CABCC/2020\|EPI_ISL_575498\|20 | ···················· | ·A·················· | ···················· | ···················· | ···················· | ···A················ |
| L | 4445 hCoV-19/England/ALDP-9D9104/2020\|EPI_ISL_576700\|20 | ···················· | ···················· | ···················· | ···················· | ···················· | ···················· |
| L | 4446 hCoV-19/England/ALDP-9D9113/2020\|EPI_ISL_576701\|20 | ···················· | ···················· | ···················· | ···················· | ···················· | ···················· |
| L | 4447 hCoV-19/England/ALDP-9D916E/2020\|EPI_ISL_576703\|20 | ···················· | ·A·················· | ···················· | ···················· | ···················· | ···A················ |
| L | 4448 hCoV-19/England/ALDP-9D919B/2020\|EPI_ISL_576705\|20 | ···················· | ···················· | ···················· | ···················· | ···················· | ···················· |
| L | 4449 hCoV-19/England/ALDP-9D91AA/2020\|EPI_ISL_576706\|20 | ···················· | ···················· | ···················· | ···················· | ···················· | ···················· |
| L | 4450 hCoV-19/England/ALDP-9D91D7/2020\|EPI_ISL_576707\|20 | ···················· | ···················· | ···················· | ···················· | ···················· | ···················· |
| L | 4451 hCoV-19/England/ALDP-9D91E6/2020\|EPI_ISL_576708\|20 | ···················· | ···················· | ···················· | ···················· | ···················· | ···················· |
| L | 4452 hCoV-19/England/ALDP-9D9210/2020\|EPI_ISL_576709\|20 | ···················· | ·A·················· | ···················· | ···················· | ···················· | ···A················ |
| L | 4453 hCoV-19/England/ALDP-9D923E/2020\|EPI_ISL_576710\|20 | ···················· | ···················· | ···················· | ···················· | ···················· | ···················· |
| L | 4454 hCoV-19/England/ALDP-9D924D/2020\|EPI_ISL_576711\|20 | ···················· | ·A·······A·········· | ···················· | ···················· | ···················· | ···················· |
| L | 4455 hCoV-19/England/ALDP-9D926B/2020\|EPI_ISL_576712\|20 | ···················· | ·A·················· | ···················· | ···················· | ···················· | ···················· |
| L | 4456 hCoV-19/England/ALDP-9D9289/2020\|EPI_ISL_576713\|20 | ···················· | ···················· | ···················· | ···················· | ···················· | ···················· |
| L | 4457 hCoV-19/England/ALDP-9D9298/2020\|EPI_ISL_576714\|20 | ···················· | ···················· | ···················· | ···················· | ···················· | ···················· |
| L | 4458 hCoV-19/England/ALDP-9D92A7/2020\|EPI_ISL_576715\|20 | ···················· | ···················· | ···················· | ···················· | ···················· | ···················· |
| L | 4459 hCoV-19/England/ALDP-9D92B6/2020\|EPI_ISL_576716\|20 | ···················· | ···················· | ···················· | ···················· | ···················· | ···················· |
| L | 4460 hCoV-19/England/ALDP-9D930E/2020\|EPI_ISL_576717\|20 | ···················· | ···················· | ···················· | ···················· | ···················· | ···················· |
| L | 4461 hCoV-19/England/ALDP-9D931D/2020\|EPI_ISL_576718\|20 | ···················· | ···················· | ···················· | ···················· | ···················· | ···················· |
| L | 4462 hCoV-19/England/ALDP-9D932C/2020\|EPI_ISL_576719\|20 | ···················· | ···················· | ···················· | ···················· | ···················· | ···················· |
| L | 4463 hCoV-19/England/ALDP-9D933B/2020\|EPI_ISL_576720\|20 | ···················· | ···················· | ···················· | ···················· | ···················· | ···················· |
| L | 4464 hCoV-19/England/ALDP-9D9377/2020\|EPI_ISL_576721\|20 | ···················· | ···················· | ···················· | ···················· | ···················· | ···················· |
| L | 4465 hCoV-19/England/ALDP-9D9386/2020\|EPI_ISL_576722\|20 | ···················· | ···················· | ···················· | ···················· | ···················· | ···················· |
| L | 4466 hCoV-19/England/ALDP-9D93E0/2020\|EPI_ISL_576724\|20 | ···················· | ···················· | ···················· | ···················· | ···················· | ···················· |
| L | 4467 hCoV-19/England/ALDP-9D93FF/2020\|EPI_ISL_576725\|20 | ···················· | ···················· | ···················· | ···················· | ···················· | ···················· |
| L | 4468 hCoV-19/England/ALDP-9D941A/2020\|EPI_ISL_576727\|20 | ···················· | ···················· | ···················· | ···················· | ···················· | ···················· |
| L | 4469 hCoV-19/England/ALDP-9D9429/2020\|EPI_ISL_576728\|20 | ···················· | ···················· | ···················· | ···················· | ···················· | ···················· |
| L | 4470 hCoV-19/England/ALDP-9D9438/2020\|EPI_ISL_576729\|20 | ···················· | ···················· | ···················· | ···················· | ···················· | ···················· |
| L | 4471 hCoV-19/England/ALDP-9D9456/2020\|EPI_ISL_576730\|20 | ···················· | ···················· | ···················· | ···················· | ···················· | ···················· |
| L | 4472 hCoV-19/England/ALDP-9D9474/2020\|EPI_ISL_576731\|20 | ···················· | ···················· | ···················· | ···················· | ···················· | ···················· |
| L | 4473 hCoV-19/England/ALDP-9D94CF/2020\|EPI_ISL_576732\|20 | ···················· | ···················· | ···················· | ···················· | ···················· | ···················· |
| L | 4474 hCoV-19/England/ALDP-9D94DE/2020\|EPI_ISL_576733\|20 | ···················· | ···················· | ···················· | ···················· | ···················· | ···················· |
| L | 4475 hCoV-19/England/ALDP-9D94ED/2020\|EPI_ISL_576734\|20 | ···················· | ···················· | ···················· | ···················· | ···················· | ···················· |
| L | 4476 hCoV-19/England/ALDP-9D9517/2020\|EPI_ISL_576735\|20 | ···················· | ···················· | ···················· | ···················· | ···················· | ···················· |
| L | 4477 hCoV-19/England/ALDP-9D9526/2020\|EPI_ISL_576736\|20 | ···················· | ···················· | ···················· | ···················· | ···················· | ···················· |
| L | 4478 hCoV-19/England/ALDP-9D9535/2020\|EPI_ISL_576737\|20 | ···················· | ·A·················· | ···················· | ···················· | ···················· | ···A················ |
| L | 4479 hCoV-19/England/ALDP-9D9562/2020\|EPI_ISL_576738\|20 | ···················· | ···················· | ···················· | ···················· | ···················· | ···················· |
| L | 4480 hCoV-19/England/ALDP-9D9571/2020\|EPI_ISL_576739\|20 | ···················· | ···················· | ···················· | ···················· | ···················· | ···················· |
| L | 4481 hCoV-19/England/ALDP-9D9580/2020\|EPI_ISL_576740\|20 | ···················· | ···················· | ···················· | ···················· | ···················· | ···················· |
| L | 4482 hCoV-19/England/ALDP-9D959F/2020\|EPI_ISL_576741\|20 | ···················· | ···················· | ···················· | ···················· | ···················· | ···················· |
| L | 4483 hCoV-19/England/ALDP-9D95AE/2020\|EPI_ISL_576742\|20 | ···················· | ···················· | ···················· | ···················· | ···················· | ···················· |
| L | 4484 hCoV-19/England/MILK-9D5478/2020\|EPI_ISL_577060\|20 | ···················· | ·A·················· | ···················· | ···················· | ···················· | ···A················ |
| L | 4485 hCoV-19/England/MILK-9D5496/2020\|EPI_ISL_577061\|20 | ···················· | ···················· | ···················· | ···················· | ···················· | ···················· |
| L | 4486 hCoV-19/England/MILK-9E12E8/2020\|EPI_ISL_577068\|20 | ···················· | ···················· | ···················· | ···················· | ···················· | ···················· |

```
L 4487 hCoV-19/England/MILK-9E12F7/2020|EPI_ISL_577069|20 ···················· ·A·················· ···················· ···················· ···················· ··A·················
L 4488 hCoV-19/England/MILK-9E1303/2020|EPI_ISL_577070|20 ···················· ···················· ···················· ···················· ···················· ····················
L 4489 hCoV-19/England/MILK-9E1330/2020|EPI_ISL_577071|20 ···················· ···················· ···················· ···················· ···················· ····················
L 4490 hCoV-19/England/MILK-9E134F/2020|EPI_ISL_577072|20 ···················· ···················· ···················· ···················· ···················· ····················
L 4491 hCoV-19/England/MILK-9E13F4/2020|EPI_ISL_577073|20 ···················· ·A·················· ···················· ···················· ···················· ····················
L 4492 hCoV-19/England/MILK-9E15B2/2020|EPI_ISL_577074|20 ···················· ···················· ···················· ···················· ···················· ····················
L 4493 hCoV-19/England/MILK-9E15C1/2020|EPI_ISL_577075|20 ···················· ···················· ···················· ···················· ···················· ····················
L 4494 hCoV-19/England/MILK-9E15D0/2020|EPI_ISL_577076|20 ···················· ···················· ···················· ···················· ···················· ····················
L 4495 hCoV-19/England/MILK-9E15FE/2020|EPI_ISL_577077|20 ···················· ···················· ···················· ···················· ···················· ····················
L 4496 hCoV-19/England/MILK-9E1628/2020|EPI_ISL_577078|20 ···················· ···················· ···················· ···················· ···················· ····················
L 4497 hCoV-19/England/MILK-9E1655/2020|EPI_ISL_577080|20 ···················· ···················· ···················· ···················· ···················· ····················
L 4498 hCoV-19/England/MILK-9E1673/2020|EPI_ISL_577081|20 ···················· ···················· ···················· ···················· ···················· ····················
L 4499 hCoV-19/England/MILK-9E16DD/2020|EPI_ISL_577082|20 ···················· ···················· ···················· ···················· ···················· ····················
L 4500 hCoV-19/England/MILK-9E1716/2020|EPI_ISL_577083|20 ···················· ···················· ···················· ···················· ···················· ····················
L 4501 hCoV-19/England/MILK-9E1725/2020|EPI_ISL_577084|20 ···················· ···················· ···················· ···················· ···················· ····················
L 4502 hCoV-19/England/MILK-9E1752/2020|EPI_ISL_577085|20 ···················· ···················· ···················· ···················· ···················· ····················
L 4503 hCoV-19/England/MILK-9E178F/2020|EPI_ISL_577086|20 ···················· ···················· ···················· ···················· ···················· ····················
L 4504 hCoV-19/England/MILK-9E17DA/2020|EPI_ISL_577087|20 ···················· ···················· ···················· ···················· ···················· ····················
L 4505 hCoV-19/England/MILK-9E17F8/2020|EPI_ISL_577088|20 ···················· ···················· ···················· ···················· ···················· ····················
L 4506 hCoV-19/England/MILK-9E1822/2020|EPI_ISL_577089|20 ···················· ···················· ···················· ···················· ······T············· ····················
L 4507 hCoV-19/England/MILK-9E1831/2020|EPI_ISL_577090|20 ···················· ···················· ···················· ···················· ···················· ····················
L 4508 hCoV-19/England/MILK-9E18E6/2020|EPI_ISL_577091|20 ···················· ···················· ···················· ···················· ······T············· ····················
L 4509 hCoV-19/England/MILK-9E18F5/2020|EPI_ISL_577092|20 ···················· ···················· ···················· ···················· ······T············· ····················
L 4510 hCoV-19/England/MILK-9E1901/2020|EPI_ISL_577093|20 ···················· ···················· ···················· ···················· ···················· ····················
L 4511 hCoV-19/England/MILK-9E1AC2/2020|EPI_ISL_577094|20 ···················· ···················· ···················· ···················· ···················· ····················
L 4512 hCoV-19/England/MILK-9E1AD1/2020|EPI_ISL_577095|20 ···················· ·A·················· ···················· ···················· ···················· ··A·················
L 4513 hCoV-19/England/NORW-F03EC/2020|EPI_ISL_577099|202 ···················· ···················· ···················· ···················· ···················· ····················
L 4514 hCoV-19/Ireland/KE-NVRL-73IRL40359/2020|EPI_ISL_57 ···················· ···················· ···················· ···················· ···················· ····················
L 4515 hCoV-19/Ireland/KK-NVRL-72IRL24802/2020|EPI_ISL_57 ···················· ···················· ···················· ···················· ···················· ····················
L 4526 hCoV-19/USA/WI-WSLH-200153/2020|EPI_ISL_578380|202 ···················· ···················· ···················· ···················· ···················· ····················
L 4527 hCoV-19/USA/WI-WSLH-200154/2020|EPI_ISL_578381|202 ···················· ···················· ···················· ···················· ···················· ····················
L 4532 hCoV-19/USA/WI-WSLH-200159/2020|EPI_ISL_578386|202 ···················· ···················· ···················· ···················· ···················· ····················
L 4533 hCoV-19/USA/WI-WSLH-200161/2020|EPI_ISL_578387|202 ···················· ···················· ···················· ···················· ···················· ····················
L 4534 hCoV-19/USA/WI-WSLH-200162/2020|EPI_ISL_578388|202 ···················· ···················· ···················· ···················· ···················· ····················
L 4543 hCoV-19/USA/WI-WSLH-200171/2020|EPI_ISL_578397|202 ···················· ···················· ···················· ···················· ···················· ····················
L 4544 hCoV-19/USA/WI-WSLH-200172/2020|EPI_ISL_578398|202 ···················· ···················· ···················· ···················· ···················· ····················
L 4546 hCoV-19/England/CAMC-9EC65A/2020|EPI_ISL_580621|20 ···················· ···················· ···················· ···················· ···················· ····················
L 4547 hCoV-19/England/ALDP-9E6EEF/2020|EPI_ISL_580622|20 ···················· ···················· ···················· ···················· ···················· ····················
L 4548 hCoV-19/England/ALDP-9EDA8A/2020|EPI_ISL_580681|20 ···················· ···················· ···················· ···················· ···················· ····················
L 4549 hCoV-19/England/ALDP-9EDAE4/2020|EPI_ISL_580682|20 ···················· ·A·················· ···················· ···················· ···················· ····················
L 4550 hCoV-19/England/MILK-9EE8DA/2020|EPI_ISL_580683|20 ···················· ···················· ···················· ···················· ···················· ····················
L 4551 hCoV-19/England/MILK-9EEEE7/2020|EPI_ISL_580685|20 ···················· ···················· ···················· ···················· ···················· ····················
L 4552 hCoV-19/England/MILK-9EE430/2020|EPI_ISL_580687|20 ···················· ···················· ···················· ···················· ···················· ····················
L 4553 hCoV-19/England/MILK-9EEDDB/2020|EPI_ISL_580688|20 ···················· ···················· ···················· ···················· ···················· ····················
L 4554 hCoV-19/England/ALDP-9EDAD5/2020|EPI_ISL_580689|20 ···················· ···················· ···················· ···················· ···················· ····················
L 4555 hCoV-19/England/ALDP-9EDC2A/2020|EPI_ISL_580690|20 ···················· ···················· ···················· ···················· ···················· ····················
L 4556 hCoV-19/England/ALDP-9EDA4E/2020|EPI_ISL_580691|20 ···················· ···················· ···················· ···················· ···················· ····················
L 4557 hCoV-19/England/ALDP-9EDC39/2020|EPI_ISL_580692|20 ···················· ···················· ···················· ···················· ···················· ····················
L 4558 hCoV-19/England/MILK-9EE870/2020|EPI_ISL_580694|20 ···················· ···················· ···················· ···················· ···················· ····················
L 4559 hCoV-19/England/ALDP-9ED8BD/2020|EPI_ISL_580695|20 ···················· ···················A ···················· ···················· ···················· ···················· 1
L 4560 hCoV-19/England/ALDP-9EDCDF/2020|EPI_ISL_580697|20 ···················· ···················· ···················· ···················· ···················· ····················
L 4561 hCoV-19/Wales/MILK-9F0565/2020|EPI_ISL_580698|2020 ···················· ···················· ···················· ···················· ···················· ····················
L 4562 hCoV-19/England/ALDP-9EDC48/2020|EPI_ISL_580699|20 ···················· ···················· ···················· ···················· ···················· ····················
L 4563 hCoV-19/England/MILK-9E30BF/2020|EPI_ISL_581253|20 ···················· ·A·················· ···················· ···················· ···················· ··A·················
L 4564 hCoV-19/Wales/MILK-9E342C/2020|EPI_ISL_581254|2020 ···················· ···················· ···················· ···················· ···················· ····················
L 4565 hCoV-19/England/MILK-9E2250/2020|EPI_ISL_581255|20 ···················· ···················· ···················· ···················· ···················· ····················
L 4566 hCoV-19/England/MILK-9E335C/2020|EPI_ISL_581256|20 ···················· ···················· ···················· ···················· ···················· ····················
L 4567 hCoV-19/England/MILK-9E2CF8/2020|EPI_ISL_581257|20 ···················· ···················· ···················· ···················· ···················· ····················
L 4568 hCoV-19/England/MILK-9E1D8D/2020|EPI_ISL_581258|20 ···················· ···················· ···················· ···················· ···················· ····················
L 4569 hCoV-19/Wales/MILK-9E2126/2020|EPI_ISL_581259|2020 ···················· ···················· ···················· ···················· ···················· ····················
L 4570 hCoV-19/England/MILK-9E2B0A/2020|EPI_ISL_581260|20 ···················· ···················· ···················· ···················· ···················· ····················
L 4571 hCoV-19/England/QEUH-9DE758/2020|EPI_ISL_581262|20 ···················· ·A·················· ···················· ···················· ···················· ··A·················
L 4572 hCoV-19/Wales/MILK-9E340E/2020|EPI_ISL_581263|2020 ···················· ···················· ···················· ···················· ···················· ····················
L 4573 hCoV-19/England/MILK-9E1EA8/2020|EPI_ISL_581264|20 ···················· ···················· ···················· ···················· ···················· ····················
L 4574 hCoV-19/England/MILK-9E1F3C/2020|EPI_ISL_581265|20 ···················· ···················· ···················· ···················· ···················· ····················
L 4575 hCoV-19/England/MILK-9E2056/2020|EPI_ISL_581266|20 ···················· ·A·················· ···················· ···················· ···················· ····················
L 4576 hCoV-19/England/QEUH-9DECA4/2020|EPI_ISL_581267|20 ···················· ···················· ···················· ···················· ····T··············· ····················
L 4577 hCoV-19/England/MILK-9E2D7D/2020|EPI_ISL_581268|20 ···················· ···················· ···················· ···················· ···················· ····················
L 4578 hCoV-19/England/MILK-9E3037/2020|EPI_ISL_581269|20 ···················· ···················· ···················· ···················· ···················· ····················
L 4579 hCoV-19/England/MILK-9E1ED5/2020|EPI_ISL_581270|20 ···················· ···················· ···················· ···················· ···················· ····················
L 4580 hCoV-19/Wales/MILK-9E1E4E/2020|EPI_ISL_581272|2020 ···················· ···················· ···················· ···················· ···················· ····················
L 4581 hCoV-19/England/MILK-9E327D/2020|EPI_ISL_581273|20 ···················· ·A·················· ···················· ···················· ···················· ··A·················
L 4582 hCoV-19/England/MILK-9E2BDD/2020|EPI_ISL_581274|20 ···················· ···················· ···················· ···················· ···················· ····················
L 4583 hCoV-19/Wales/MILK-9E2B28/2020|EPI_ISL_581275|2020 ···················· ···················· ···················· ···················· ···················· ····················
L 4584 hCoV-19/England/MILK-9E3028/2020|EPI_ISL_581276|20 ···················· ···················· ···················· ···················· ···················· ····················
L 4585 hCoV-19/England/MILK-9E2E10/2020|EPI_ISL_581277|20 ···················· ···················· ···················· ···················· ···················· ····················
L 4586 hCoV-19/Wales/MILK-9E2A2B/2020|EPI_ISL_581278|2020 ···················· ···················· ···················· ···················· ···················· ····················
L 4587 hCoV-19/England/MILK-9E2B91/2020|EPI_ISL_581279|20 ···················· ···················· ···················· ···················· ···················· ····················
L 4588 hCoV-19/England/MILK-9E31E9/2020|EPI_ISL_581280|20 ···················· ···················· ···················· ···················· ···················· ····················
L 4589 hCoV-19/England/MILK-9E2205/2020|EPI_ISL_581281|20 ···················· ···················· ···················· ···················· ···················· ····················
```

L 4590 hCoV-19/Wales/MILK-9E2E2F/2020|EPI_ISL_581282|2020 ···················· ···················· ···················· ···················· ···················· ····················
L 4591 hCoV-19/Wales/MILK-9E20CF/2020|EPI_ISL_581283|2020 ···················· ···················· ···················· ···················· ···················· ····················
L 4592 hCoV-19/Wales/MILK-9E32B9/2020|EPI_ISL_581284|2020 ···················· ···················· ···················· ···················· ···················· ····················
L 4593 hCoV-19/England/QEUH-9DE5C7/2020|EPI_ISL_581285|20 ···················· ·A·················· ···················· ···················· ···················· ···A················
L 4594 hCoV-19/Wales/MILK-9E32AA/2020|EPI_ISL_581287|2020 ···················· ···················· ···················· ···················· ···················· ····················
L 4595 hCoV-19/England/MILK-9E3486/2020|EPI_ISL_581288|20 ···················· ···················· ···················· ···················· ···················· ····················
L 4596 hCoV-19/England/MILK-9E1D6F/2020|EPI_ISL_581289|20 ···················· ···················· ···················· ···················· ···················· ····················
L 4597 hCoV-19/England/MILK-9E2E4D/2020|EPI_ISL_581290|20 ···················· ·A·················· ···················· ···················· ···················· ···A················
L 4598 hCoV-19/England/QEUH-9DEF05/2020|EPI_ISL_581291|20 ···················· ···················· ···················· ···················· ···················· ····················
L 4599 hCoV-19/England/MILK-9E3398/2020|EPI_ISL_581292|20 ···················· ·A·······A·········· ···················· ···················· ···················· ····················
L 4600 hCoV-19/England/MILK-9E1F69/2020|EPI_ISL_581293|20 ···················· ···················· ···················· ···················· ······T············· ····················
L 4601 hCoV-19/England/MILK-9E2DE6/2020|EPI_ISL_581294|20 ···················· ···················· ···················· ···················· ···················· ····················
L 4602 hCoV-19/England/MILK-9E2092/2020|EPI_ISL_581295|20 ···················· ···················· ···················· ···················· ···················· ····················
L 4603 hCoV-19/England/MILK-9E2AC1/2020|EPI_ISL_581296|20 ···················· ···················· ···················· ···················· ···················· ····················
L 4604 hCoV-19/England/MILK-9E1C9F/2020|EPI_ISL_581297|20 ···················· ···················· ···················· ···················· ···················· ····················
L 4605 hCoV-19/England/MILK-9E3310/2020|EPI_ISL_581298|20 ···················· ···················· ···················· ···················· ···················· ····················
L 4606 hCoV-19/England/MILK-9E3301/2020|EPI_ISL_581299|20 ···················· ···················· ···················· ···················· ···················· ····················
L 4609 hCoV-19/USA/VA-DCLS-1350/2020|EPI_ISL_572266|2020- ···················· ···················· ···················· ···················· ···················· ····················
L 4610 hCoV-19/USA/VA-DCLS-1354/2020|EPI_ISL_572267|2020- ···················· ···················· ···················· ···················· ···················· ····················
L 4611 hCoV-19/USA/VA-DCLS-1360/2020|EPI_ISL_572268|2020- ···················· ···················· ···················· ···················· ···················· ····················
L 4612 hCoV-19/USA/VA-DCLS-1707/2020|EPI_ISL_572269|2020- ···················· ···················· ···················· ···················· ···················· ····················
L 4613 hCoV-19/USA/VA-DCLS-1709/2020|EPI_ISL_572270|2020- ···················· ···················· ···················· ···················· ···················· ····················
L 4614 hCoV-19/USA/VA-DCLS-1190/2020|EPI_ISL_572271|2020- ···················· ···················· ···················· ···················· ···················· ····················
L 4615 hCoV-19/USA/VA-DCLS-1191/2020|EPI_ISL_572272|2020- ···················· ···················· ···················· ···················· ···················· ····················
L 4616 hCoV-19/USA/VA-DCLS-1193/2020|EPI_ISL_572273|2020- ···················· ···················· ···················· ···················· ···················· ····················
L 4617 hCoV-19/USA/VA-DCLS-1195/2020|EPI_ISL_572274|2020- ···················· ···················· ···················· ···················· ···················· ····················
L 4618 hCoV-19/USA/VA-DCLS-1197/2020|EPI_ISL_572275|2020- ···················· ···················· ···················· ···················· ···················· ····················
L 4619 hCoV-19/USA/VA-DCLS-1200/2020|EPI_ISL_572276|2020- ···················· ···················· ···················· ···················· ···················· ····················
L 4620 hCoV-19/USA/VA-DCLS-1203/2020|EPI_ISL_572277|2020- ···················· ···················· ···················· ···················· ···················· ····················
L 4621 hCoV-19/USA/VA-DCLS-1205/2020|EPI_ISL_572278|2020- ···················· ···················· ···················· ···················· ···················· ····················
L 4622 hCoV-19/USA/VA-DCLS-1208/2020|EPI_ISL_572279|2020- ···················· ···················· ···················· ···················· ···················· ·········A··········
L 4623 hCoV-19/USA/VA-DCLS-1209/2020|EPI_ISL_572280|2020- ···················· ···················· ···················· ···················· ···················· ····················
L 4624 hCoV-19/USA/VA-DCLS-1212/2020|EPI_ISL_572281|2020- ···················· ···················· ···················· ···················· ···················· ····················
L 4625 hCoV-19/USA/VA-DCLS-1213/2020|EPI_ISL_572282|2020- ···················· ···················· ···················· ···················· ···················· ····················
L 4626 hCoV-19/USA/VA-DCLS-1216/2020|EPI_ISL_572283|2020- ···················· ···················· ···················· ···················· ···················· ····················
L 4627 hCoV-19/USA/VA-DCLS-1217/2020|EPI_ISL_572284|2020- ···················· ···················· ···················· ···················· ···················· ····················
L 4628 hCoV-19/USA/VA-DCLS-1290/2020|EPI_ISL_572285|2020- ···················· ···················· ···················· ···················· ···················· ····················
L 4629 hCoV-19/USA/VA-DCLS-1294/2020|EPI_ISL_572287|2020- ···················· ···················· ···················· ···················· ···················· ····················
L 4630 hCoV-19/USA/VA-DCLS-1295/2020|EPI_ISL_572288|2020- ···················· ···················· ···················· ···················· ···················· ····················
L 4631 hCoV-19/USA/VA-DCLS-1298/2020|EPI_ISL_572289|2020- ···················· ·A·················· ···················· ···················· ···················· ····················
L 4632 hCoV-19/USA/VA-DCLS-1300/2020|EPI_ISL_572290|2020- ···················· ···················· ···················· ···················· ···················· ····················
L 4634 hCoV-19/USA/VA-DCLS-1276/2020|EPI_ISL_572292|2020- ···················· ···················· ···················· ···················· ···················· ····················
L 4635 hCoV-19/USA/VA-DCLS-1277/2020|EPI_ISL_572293|2020- ···················· ···················· ···················· ···················· ···················· ····················
L 4636 hCoV-19/USA/VA-DCLS-1279/2020|EPI_ISL_572294|2020- ···················· ···················· ···················· ···················· ···················· ····················
L 4637 hCoV-19/USA/VA-DCLS-1281/2020|EPI_ISL_572295|2020- ···················· ···················· ···················· ···················· ···················· ····················
L 4638 hCoV-19/USA/VA-DCLS-1282/2020|EPI_ISL_572296|2020- ···················· ···················· ···················· ···················· ···················· ····················
L 4639 hCoV-19/USA/VA-DCLS-1283/2020|EPI_ISL_572297|2020- ···················· ···················· ···················· ···················· ···················· ····················
L 4640 hCoV-19/USA/VA-DCLS-1284/2020|EPI_ISL_572298|2020- ···················· ···················· ···················· ···················· ···················· ····················
L 4641 hCoV-19/USA/VA-DCLS-1285/2020|EPI_ISL_572299|2020- ···················· ···················· ···················· ···················· ···················· ····················
L 4642 hCoV-19/England/QEUH-9BA6EC/2020|EPI_ISL_573597|20 ···················· ·A·················· ···················· ···················· ···················· ····················
L 4643 hCoV-19/England/QEUH-9BA725/2020|EPI_ISL_573598|20 ···················· ···················· ···················· ···················· ····T··············· ····················
L 4644 hCoV-19/England/QEUH-9BA743/2020|EPI_ISL_573599|20 ···················· ·A·················· ···················· ···················· ···················· ····················
L 4645 hCoV-19/England/MILK-9918B8/2020|EPI_ISL_577000|20 ···················· ···················· ···················· ···················· ···················· ····················
L 4646 hCoV-19/England/MILK-9918E5/2020|EPI_ISL_577001|20 ···················· ···················· ···················· ···················· ···················· ····················
L 4647 hCoV-19/England/MILK-9918F4/2020|EPI_ISL_577002|20 ···················· ······C············· ···················· ···················· ···················· ····················
L 4648 hCoV-19/England/MILK-99192E/2020|EPI_ISL_577003|20 ···················· ·A·················· ···················· ···················· ···················· ····················
L 4649 hCoV-19/England/MILK-99193D/2020|EPI_ISL_577004|20 ···················· ···················· ···················· ···················· ···················· ····················
L 4650 hCoV-19/England/MILK-991979/2020|EPI_ISL_577006|20 ···················· ·A·······A·········· ···················· ···················· ···················· ····················
L 4651 hCoV-19/England/MILK-991988/2020|EPI_ISL_577007|20 ···················· ···················· ···················· ···················· ···················· ····················
L 4653 hCoV-19/England/MILK-9919D3/2020|EPI_ISL_577011|20 ···················· ···················· ···················· ···················· ···················· ····················
L 4655 hCoV-19/England/MILK-991A1C/2020|EPI_ISL_577013|20 ···················· ···················· ···················· ···················· ···················· ····················
L 4657 hCoV-19/England/MILK-991A58/2020|EPI_ISL_577016|20 ···················· ···················· ···················· ···················· ···················· ····················
L 4658 hCoV-19/England/MILK-991A76/2020|EPI_ISL_577017|20 ···················· ···················· ···················· ···················· ···················· ····················
L 4659 hCoV-19/England/MILK-991AD0/2020|EPI_ISL_577019|20 ···················· ···················· ···················· ···················· ···················· ····················
L 4660 hCoV-19/England/MILK-991AEF/2020|EPI_ISL_577020|20 ···················· ···················· ···················· ···················· ···················· ····················
L 4661 hCoV-19/England/MILK-991AFE/2020|EPI_ISL_577021|20 ···················· ···················· ···················· ···················· ···················· ····················
L 4662 hCoV-19/England/MILK-991B19/2020|EPI_ISL_577022|20 ···················· ···················· ···················· ···················· ···················· ····················
L 4663 hCoV-19/England/MILK-991B28/2020|EPI_ISL_577023|20 ···················· ···················· ···················· ···················· ···················· ····················
L 4664 hCoV-19/England/MILK-991B37/2020|EPI_ISL_577024|20 ···················· ···················· ···················· ···················· ···················· ····················
L 4665 hCoV-19/England/MILK-991B46/2020|EPI_ISL_577025|20 ···················· ···················· ···················· ···················· ···················· ····················
L 4666 hCoV-19/England/MILK-991B64/2020|EPI_ISL_577026|20 ··················T· ···················· ···················· ···················· ······T············· ···················· 1
L 4667 hCoV-19/England/MILK-991B82/2020|EPI_ISL_577027|20 ···················· ·A·················· ···················· ···················· ···················· ···A················
L 4668 hCoV-19/England/MILK-991B91/2020|EPI_ISL_577028|20 ···················· ···················· ···················· ···················· ···················· ····················
L 4669 hCoV-19/England/MILK-991BBF/2020|EPI_ISL_577029|20 ···················· ···················· ···················· ···················· ···················· ····················
L 4670 hCoV-19/England/MILK-991BEC/2020|EPI_ISL_577030|20 ···················· ···················· ···················· ···················· ···················· ····················
L 4672 hCoV-19/England/MILK-991C07/2020|EPI_ISL_577032|20 ···················· ···················· ···················· ···················· ···················· ····················
L 4673 hCoV-19/England/MILK-991C16/2020|EPI_ISL_577033|20 ···················· ···················· ···················· ···················· ···················· ····················
L 4674 hCoV-19/England/MILK-991C25/2020|EPI_ISL_577034|20 ···················· ···················· ···················· ···················· ···················· ····················
L 4675 hCoV-19/England/MILK-991C52/2020|EPI_ISL_577035|20 ···················· ···················· ···················· ···················· ···················· ····················
L 4677 hCoV-19/England/MILK-991CDA/2020|EPI_ISL_577038|20 ···················· ···················· ···················· ···················· ······T············· ····················

| | | | | | | | |
|---|---|---|---|---|---|---|---|
| L | 4678 hCoV-19/Ireland/KE-NVRL-73IRL40299/2020\|EPI_ISL_57 | ···················· | ···················· | ···················· | ···················· | ···················· | ···················· |
| L | 4679 hCoV-19/Ireland/KK-NVRL-72IRL32188/2020\|EPI_ISL_57 | ···················· | ···················· | ···················· | ···················· | ···················· | ···················· |
| L | 4680 hCoV-19/Ireland/KK-NVRL-72IRL37138/2020\|EPI_ISL_57 | ···················· | ···················· | ···················· | ···················· | ···················· | ···················· |
| L | 4681 hCoV-19/Ireland/KK-NVRL-72IRL37173/2020\|EPI_ISL_57 | ···················· | ···················· | ···················· | ···················· | ···················· | ···················· |
| L | 4682 hCoV-19/Ireland/KK-NVRL-72IRL40595/2020\|EPI_ISL_57 | ···················· | ·A·················· | ···················· | ···················· | ···················· | ···A················ |
| L | 4683 hCoV-19/Ireland/KK-NVRL-72IRL50388/2020\|EPI_ISL_57 | ···················· | ···················· | ···················· | ···················· | ···················· | ···················· |
| L | 4685 hCoV-19/Ireland/LH-NVRL-73IRL27298/2020\|EPI_ISL_57 | ···················· | ···················· | ···················· | ···················· | ···················· | ···················· |
| L | 4686 hCoV-19/Ireland/LK-NVRL-72IRL34726/2020\|EPI_ISL_57 | ···················· | ·A·················· | ···················· | ···················· | ···················· | ···A················ |
| L | 4687 hCoV-19/Ireland/LK-NVRL-72IRL77937/2020\|EPI_ISL_57 | ···················· | ·····C·············· | ···················· | ···················· | ···················· | ···················· |
| L | 4688 hCoV-19/Ireland/LK-NVRL-73IRL25283/2020\|EPI_ISL_57 | ···················· | ·A·················· | ···················· | ···················· | ···················· | ···A················ |
| L | 4689 hCoV-19/Ireland/LK-NVRL-73IRL26070/2020\|EPI_ISL_57 | ···················· | ···················· | ···················· | ···················· | ···················· | ···················· |
| L | 4690 hCoV-19/Ireland/LK-NVRL-73IRL28138/2020\|EPI_ISL_57 | ···················· | ·····C·············· | ···················· | ···················· | ···················· | ···················· |
| L | 4691 hCoV-19/Ireland/LK-NVRL-73IRL28235/2020\|EPI_ISL_57 | ···················· | ·A·················· | ···················· | ···················· | ···················· | ···A················ |
| L | 4692 hCoV-19/Ireland/MH-NVRL-73IRL26442/2020\|EPI_ISL_57 | ···················· | ···················· | ···················· | ···················· | ···················· | ···················· |
| L | 4693 hCoV-19/Ireland/OY-NVRL-72IRL51564/2020\|EPI_ISL_57 | ···················· | ···················· | ···················· | ···················· | ···················· | ···················· |
| L | 4694 hCoV-19/Ireland/OY-NVRL-73IRL30167/2020\|EPI_ISL_57 | ···················· | ···················· | ···················· | ···················· | ···················· | ···················· |
| L | 4695 hCoV-19/Ireland/RN-NVRL-73IRL30636/2020\|EPI_ISL_57 | ···················· | ···················· | ···················· | ···················· | ···················· | ···················· |
| L | 4696 hCoV-19/Ireland/SO-NVRL-73IRL13569/2020\|EPI_ISL_57 | ···················· | ···················· | ···················· | ···················· | ···················· | ···················· |
| L | 4697 hCoV-19/Ireland/WD-NVRL-20ILR05743/2020\|EPI_ISL_57 | ···················· | ·A·················· | ···················· | ···················· | ···················· | ···················· |
| L | 4698 hCoV-19/Ireland/WD-NVRL-72IRL32318/2020\|EPI_ISL_57 | ···················· | ···················· | ···················· | ···················· | ···················· | ···················· |
| L | 4699 hCoV-19/Ireland/WD-NVRL-72IRL32320/2020\|EPI_ISL_57 | ···················· | ·A·················· | ···················· | ···················· | ···················· | ···················· |
| L | 4702 hCoV-19/Ireland/WD-NVRL-72IRL52747/2020\|EPI_ISL_57 | ···················· | ·A·················· | ···················· | ···················· | ···················· | ···················· |
| L | 4703 hCoV-19/Ireland/WD-NVRL-72IRL52753/2020\|EPI_ISL_57 | ···················· | ·A·················· | ···················· | ···················· | ···················· | ···················· |
| L | 4704 hCoV-19/Ireland/WW-NVRL-73IRL23909/2020\|EPI_ISL_57 | ···················· | ···················· | ···················· | ···················· | ···················· | ···················· |
| L | 4706 hCoV-19/Ireland/WW-NVRL-73IRL23933/2020\|EPI_ISL_57 | ···················· | ···················· | ···················· | ···················· | ···················· | ···················· |
| L | 4708 hCoV-19/Ireland/WW-NVRL-73IRL29985/2020\|EPI_ISL_57 | ···················· | ···················· | ···················· | ···················· | ···················· | ···················· |
| L | 4709 hCoV-19/Ireland/WX-NVRL-72IRL33055/2020\|EPI_ISL_57 | ···················· | ···················· | ···················· | ···················· | ···················· | ···················· |
| L | 4710 hCoV-19/Ireland/WX-NVRL-72IRL33081/2020\|EPI_ISL_57 | ···················· | ·A·················· | ···················· | ···················· | ···················· | ···A················ |
| L | 4711 hCoV-19/USA/WI-WSLH-200103/2020\|EPI_ISL_578335\|202 | ···················· | ···················· | ···················· | ···················· | ···················· | ···················· |
| L | 4712 hCoV-19/USA/WI-WSLH-200104/2020\|EPI_ISL_578336\|202 | ···················· | ···················· | ···················· | ···················· | ···················· | ···················· |
| L | 4714 hCoV-19/USA/WI-WSLH-200107/2020\|EPI_ISL_578338\|202 | ···················· | ···················· | ···················· | ···················· | ···················· | ···················· |
| L | 4715 hCoV-19/USA/WI-WSLH-200108/2020\|EPI_ISL_578339\|202 | ···················· | ···················· | ···················· | ···················· | ···················· | ···················· |
| L | 4717 hCoV-19/USA/WI-WSLH-200110/2020\|EPI_ISL_578341\|202 | ···················· | ···················· | ···················· | ···················· | ···················· | ···················· |
| L | 4718 hCoV-19/USA/WI-WSLH-200111/2020\|EPI_ISL_578342\|202 | ···················· | ···················· | ···················· | ···················· | ···················· | ···················· |
| L | 4720 hCoV-19/USA/WI-WSLH-200113/2020\|EPI_ISL_578344\|202 | ···················· | ···················· | ···················· | ···················· | ···················· | ···················· |
| L | 4724 hCoV-19/USA/WI-WSLH-200118/2020\|EPI_ISL_578348\|202 | ···················· | ···················· | ···················· | ···················· | ···················· | ···················· |
| L | 4725 hCoV-19/USA/WI-WSLH-200119/2020\|EPI_ISL_578349\|202 | ···················· | ···················· | ···················· | ···················· | ···················· | ···················· |
| L | 4730 hCoV-19/USA/WI-WSLH-200125/2020\|EPI_ISL_578354\|202 | ···················· | ···················· | ···················· | ···················· | ···················· | ···················· |
| L | 4737 hCoV-19/USA/WI-WSLH-200133/2020\|EPI_ISL_578361\|202 | ···················· | ···················· | ···················· | ···················· | ···················· | ···················· |
| L | 4746 hCoV-19/Canada/NS-NML-1219/2020\|EPI_ISL_579616\|202 | ···················· | ···················· | ···················· | ···················· | ···················· | ···················· |
| L | 4747 hCoV-19/Canada/NS-NML-1223/2020\|EPI_ISL_579619\|202 | ···················· | ···················· | ···················· | ···················· | ···················· | ···················· |
| L | 4748 hCoV-19/Canada/NS-NML-1244/2020\|EPI_ISL_579636\|202 | ···················· | ···················· | ···················· | ···················· | ···················· | ···················· |
| L | 4749 hCoV-19/Canada/NS-NML-1246/2020\|EPI_ISL_579638\|202 | ···················· | ···················· | ···················· | ···················· | ···················· | ···················· |
| L | 4750 hCoV-19/Canada/NS-NML-1249/2020\|EPI_ISL_579640\|202 | ···················· | ···················· | ···················· | ···················· | ···················· | ···················· |
| L | 4751 hCoV-19/Canada/NS-NML-1261/2020\|EPI_ISL_579649\|202 | ···················· | ···················· | ···················· | ···················· | ···················· | ···················· |
| L | 4752 hCoV-19/Canada/NS-NML-1277/2020\|EPI_ISL_579664\|202 | ···················· | ···················· | ···················· | ···················· | ···················· | ···················· |
| L | 4753 hCoV-19/Canada/NS-NML-1285/2020\|EPI_ISL_579669\|202 | ···················· | ···················· | ···················· | ···················· | ···················· | ···················· |
| L | 4754 hCoV-19/Canada/NS-NML-1296/2020\|EPI_ISL_579679\|202 | ···················· | ···················· | ···················· | ···················· | ···················· | ···················· |
| L | 4755 hCoV-19/Canada/NS-NML-1316/2020\|EPI_ISL_579696\|202 | ···················· | ···················· | ···················· | ···················· | ···················· | ···················· |
| L | 4756 hCoV-19/England/ALDP-9E71B8/2020\|EPI_ISL_580600\|20 | ···················· | ···················· | ···················· | ···················· | ···················· | ···················· |
| L | 4757 hCoV-19/England/ALDP-9E6C12/2020\|EPI_ISL_580601\|20 | ···················· | ···················· | ···················· | ···················· | ···················· | ···················· |
| L | 4758 hCoV-19/England/ALDP-9E6526/2020\|EPI_ISL_580602\|20 | ···················· | ···················· | ···················· | ···················· | ···················· | ···················· |
| L | 4759 hCoV-19/England/CAMC-9EC62D/2020\|EPI_ISL_580603\|20 | ···················· | ···················· | ···················· | ···················· | ···················· | ···················· |
| L | 4760 hCoV-19/England/ALDP-9E6D4C/2020\|EPI_ISL_580604\|20 | ···················· | ···················· | ···················· | ···················· | ···················· | ···················· |
| L | 4761 hCoV-19/England/ALDP-9E72A6/2020\|EPI_ISL_580605\|20 | ···················· | ···················· | ···················· | ···················· | ···················· | ···················· |
| L | 4762 hCoV-19/England/ALDP-9E7200/2020\|EPI_ISL_580606\|20 | ···················· | ···················· | ···················· | ···················· | ···················· | ···················· |
| L | 4763 hCoV-19/England/CAMC-9EC46F/2020\|EPI_ISL_580607\|20 | ···················· | ···················· | ···················· | ···················· | ···················· | ···················· |
| L | 4764 hCoV-19/England/ALDP-9E6F64/2020\|EPI_ISL_580608\|20 | ···················· | ···················· | ···················· | ···················· | ···················· | ···················· |
| L | 4765 hCoV-19/England/CAMC-9EC405/2020\|EPI_ISL_580610\|20 | ···················· | ···················· | ···················· | ···················· | ···················· | ···················· |
| L | 4766 hCoV-19/England/ALDP-9E6EB2/2020\|EPI_ISL_580611\|20 | ···················· | ···················· | ···················· | ···················· | ···················· | ···················· |
| L | 4767 hCoV-19/England/ALDP-9E675D/2020\|EPI_ISL_580612\|20 | ···················· | ·A·················· | ···················· | ···················· | ···················· | ···A················ |
| L | 4768 hCoV-19/England/ALDP-9E69B1/2020\|EPI_ISL_580613\|20 | ···················· | ···················· | ···················· | ···················· | ···················· | ···················· |
| L | 4769 hCoV-19/England/ALDP-9E7051/2020\|EPI_ISL_580614\|20 | ···················· | ···················· | ···················· | ···················· | ···················· | ···················· |
| L | 4770 hCoV-19/England/ALDP-9E674E/2020\|EPI_ISL_580615\|20 | ···················· | ···················· | ···················· | ···················· | ···················· | ···················· |
| L | 4771 hCoV-19/England/ALDP-9E6D2E/2020\|EPI_ISL_580616\|20 | ···················· | ···················· | ···················· | ···················· | ···················· | ···················· |
| L | 4772 hCoV-19/England/ALDP-9E6CC7/2020\|EPI_ISL_580617\|20 | ···················· | ···················· | ···················· | ···················· | ···················· | ···················· |
| L | 4773 hCoV-19/England/CAMC-9EC308/2020\|EPI_ISL_580618\|20 | ···················· | ···················· | ···················· | ···················· | ···················· | ···················· |
| L | 4774 hCoV-19/England/ALDP-9E63A4/2020\|EPI_ISL_580620\|20 | ···················· | ···················· | ···················· | ···················· | ···················· | ···················· |
| L | 4775 hCoV-19/England/ALDP-9E6A90/2020\|EPI_ISL_580623\|20 | ···················· | ···················· | ···················· | ···················· | ···················· | ···················· |
| L | 4776 hCoV-19/England/ALDP-9E6AAF/2020\|EPI_ISL_580624\|20 | ···················· | ···················· | ···················· | ···················· | ···················· | ···················· |
| L | 4777 hCoV-19/England/CAMC-9EC450/2020\|EPI_ISL_580625\|20 | ···················· | ···················· | ···················· | ···················· | ···················· | ···················· |
| L | 4778 hCoV-19/England/CAMC-9EC186/2020\|EPI_ISL_580626\|20 | ···················· | ···················· | ···················· | ···················· | ···················· | ···················· |
| L | 4779 hCoV-19/England/ALDP-9E6F91/2020\|EPI_ISL_580627\|20 | ···················· | ·A·················· | ···················· | ···················· | ···················· | ···A················ |
| L | 4780 hCoV-19/England/ALDP-9E6EC1/2020\|EPI_ISL_580628\|20 | ···················· | ·A·················· | ···················· | ···················· | ···················· | ···················· |
| L | 4781 hCoV-19/England/ALDP-9E60DA/2020\|EPI_ISL_580629\|20 | ···················· | ···················· | ···················· | ···················· | ···················· | ···················· |
| L | 4782 hCoV-19/England/ALDP-9E6A81/2020\|EPI_ISL_580630\|20 | ···················· | ···················· | ···················· | ···················· | ···················· | ···················· |
| L | 4783 hCoV-19/England/MILK-9F0486/2020\|EPI_ISL_580631\|20 | ···················· | ···················· | ···················· | ···················· | ···················· | ···················· |
| L | 4784 hCoV-19/England/ALDP-9EDB3C/2020\|EPI_ISL_580633\|20 | ···················· | ···················· | ···················· | ···················· | ···················· | ···················· |
| L | 4785 hCoV-19/England/MILK-9F03F2/2020\|EPI_ISL_580634\|20 | ···················· | ···················· | ···················· | ···················· | ···················· | ···················· |
| L | 4786 hCoV-19/England/ALDP-9ED792/2020\|EPI_ISL_580635\|20 | ···················· | ·············A······ | ···················· | ···················· | ···················· | ···················· |

| | | | | | | | | | |
|---|---|---|---|---|---|---|---|---|---|
| L | 4787 | hCoV-19/Wales/MILK-9EE99B/2020\|EPI_ISL_580637\|2020 | ···················· | ···················· | ····················· | ···················· | ···················· | ···················· | |
| L | 4788 | hCoV-19/England/MILK-9EED26/2020\|EPI_ISL_580638\|20 | ···················· | ···················· | ····················· | ···················· | ···················· | ···················· | |
| L | 4789 | hCoV-19/England/MILK-9F041D/2020\|EPI_ISL_580639\|20 | ···················· | ···················· | ····················· | ···················· | ···················· | ···················· | |
| L | 4790 | hCoV-19/England/ALDP-9EDBB4/2020\|EPI_ISL_580640\|20 | ···················· | ···················· | ····················· | ···················· | ···················· | ···················· | |
| L | 4791 | hCoV-19/England/MILK-9EE89E/2020\|EPI_ISL_580641\|20 | ···················· | ···················· | ····················· | ···················· | ···················· | ···················· | |
| L | 4792 | hCoV-19/England/MILK-9EE53D/2020\|EPI_ISL_580642\|20 | ···················· | ···················· | ····················· | ···················· | ···················· | ···················· | |
| L | 4793 | hCoV-19/England/MILK-9EEA7A/2020\|EPI_ISL_580644\|20 | ···················· | ···················· | ····················· | ···················· | ······T············· | ···················· | |
| L | 4794 | hCoV-19/England/ALDP-9ED9BA/2020\|EPI_ISL_580645\|20 | ···················· | ···················· | ····················· | ···················· | ···················· | ···················· | |
| L | 4795 | hCoV-19/Wales/MILK-9EEB1D/2020\|EPI_ISL_580647\|2020 | ···················· | ···················· | ····················· | ···················· | ···················· | ···················· | |
| L | 4796 | hCoV-19/England/ALDP-9EDCC0/2020\|EPI_ISL_580648\|20 | ···················· | ···················· | ····················· | ···················· | ···················· | ···················· | |
| L | 4797 | hCoV-19/England/MILK-9EE816/2020\|EPI_ISL_580649\|20 | ···················· | ···················· | ····················· | ···················· | ···················· | ···················· | |
| L | 4798 | hCoV-19/England/ALDP-9EDBD2/2020\|EPI_ISL_580650\|20 | ···················· | ···················· | ····················· | ···················· | ···················· | ···················· | |
| L | 4799 | hCoV-19/England/ALDP-9EDE06/2020\|EPI_ISL_580651\|20 | ···················· | ···················· | ····················· | ···················· | ···················· | ···················· | |
| L | 4800 | hCoV-19/England/MILK-9EE852/2020\|EPI_ISL_580652\|20 | ···················· | ···················· | ····················· | ···················· | ···················· | ···················· | |
| L | 4801 | hCoV-19/England/MILK-9F036B/2020\|EPI_ISL_580653\|20 | ···················· | ···················· | ····················· | ···················· | ······T············· | ···················· | |
| L | 4802 | hCoV-19/England/ALDP-9ED941/2020\|EPI_ISL_580655\|20 | ···················· | ···················· | ····················· | ···················· | ···················· | ···················· | |
| L | 4803 | hCoV-19/Wales/MILK-9F02D7/2020\|EPI_ISL_580656\|2020 | ···················· | ···················· | ····················· | ···················· | ···················· | ···················· | |
| L | 4804 | hCoV-19/England/MILK-9F0398/2020\|EPI_ISL_580657\|20 | ···················· | ···················· | ····················· | ···················· | ···················· | ···················· | |
| L | 4805 | hCoV-19/England/MILK-9EE5D3/2020\|EPI_ISL_580658\|20 | ···················· | ···················· | ····················· | ···················· | ···················· | ···················· | |
| L | 4806 | hCoV-19/England/ALDP-9ED70B/2020\|EPI_ISL_580660\|20 | ···················· | ···················· | ···········T········· | ···················· | ···················· | ···················· | |
| L | 4807 | hCoV-19/England/MILK-9F0732/2020\|EPI_ISL_580661\|20 | ···················· | ······C············· | ····················· | ···················· | ···················· | ···················· | |
| L | 4808 | hCoV-19/England/MILK-9EEC83/2020\|EPI_ISL_580662\|20 | ···················· | ···················· | ····················· | ···················· | ···················· | ···················· | |
| L | 4809 | hCoV-19/England/ALDP-9ED738/2020\|EPI_ISL_580663\|20 | ···················· | ···················· | ····················· | ···················· | ···················· | ···················· | |
| L | 4810 | hCoV-19/England/MILK-9F0811/2020\|EPI_ISL_580665\|20 | ···················· | ···················· | ····················· | ···················· | ···················· | ···················· | |
| L | 4811 | hCoV-19/Wales/MILK-9EEBFF/2020\|EPI_ISL_580666\|2020 | ···················· | ···················· | ····················· | ···················· | ···················· | ···················· | |
| L | 4812 | hCoV-19/England/MILK-9F03A7/2020\|EPI_ISL_580667\|20 | ···················· | ···················· | ····················· | ···················· | ···················· | ···················· | |
| L | 4813 | hCoV-19/England/MILK-9EE44F/2020\|EPI_ISL_580668\|20 | ···················· | ···················· | ····················· | ···················· | ···················· | ···················· | |
| L | 4814 | hCoV-19/England/MILK-9EE5E2/2020\|EPI_ISL_580669\|20 | ···················· | ···················· | ····················· | ···················· | ···················· | ···················· | |
| L | 4815 | hCoV-19/England/MILK-9EE8F8/2020\|EPI_ISL_580670\|20 | ···················· | ···················· | ····················· | ···················· | ···················· | ···················· | |
| L | 4816 | hCoV-19/Wales/MILK-9F01E9/2020\|EPI_ISL_580671\|2020 | ···················· | ·A·················· | ····················· | ···················· | ···················· | ···A················ | |
| L | 4817 | hCoV-19/England/MILK-9EE8BC/2020\|EPI_ISL_580672\|20 | ···················· | ···················· | ····················· | ···················· | ···················· | ···················· | |
| L | 4818 | hCoV-19/England/MILK-9EEAA7/2020\|EPI_ISL_580673\|20 | ···················· | ···················· | ····················· | ···················· | ···················· | ···················· | |
| L | 4819 | hCoV-19/England/ALDP-9ED6E0/2020\|EPI_ISL_580674\|20 | ···················· | ···················· | ····················· | ···················· | ···················· | ···················· | |
| L | 4820 | hCoV-19/England/ALDP-9EDA11/2020\|EPI_ISL_580675\|20 | ···················· | ···················· | ····················· | ···················· | ···················· | ···················· | |
| L | 4821 | hCoV-19/England/ALDP-9EDB2D/2020\|EPI_ISL_580679\|20 | ···················· | ···················· | ····················· | ···················· | ···················· | ···················· | |
| L | 4822 | hCoV-19/England/ALDP-9ED9F6/2020\|EPI_ISL_580680\|20 | ···················· | ···················· | ····················· | ···················· | ···················· | ···················· | |
| L | 4823 | hCoV-19/England/ALDP-9EDC0C/2020\|EPI_ISL_580706\|20 | ···················· | ···················· | ····················· | ···················· | ···················· | ···················· | |
| L | 4824 | hCoV-19/Wales/MILK-9F0714/2020\|EPI_ISL_580707\|2020 | ···················· | ···················· | ····················· | ···················· | ···················· | ···················· | |
| L | 4826 | hCoV-19/Switzerland/BS-42374720/2020\|EPI_ISL_58193 | ···················· | ···················· | ····················· | ···················· | ·NN················· | ···················· | |
| L | 4839 | hCoV-19/Switzerland/BS-42387582/2020\|EPI_ISL_58194 | ···················· | ···················· | ····················· | ···················· | ·NN················· | ···················· | |
| L | 4841 | hCoV-19/Switzerland/BS-42389639/2020\|EPI_ISL_58194 | ···················· | ···················· | ····················· | ···················· | ·NN················· | ···················· | |
| L | 4845 | hCoV-19/Switzerland/BS-42391102/2020\|EPI_ISL_58195 | ···················· | ···················· | ····················· | ···················· | ···················· | ···················· | |
| L | 4847 | hCoV-19/Switzerland/BS-42395137/2020\|EPI_ISL_58195 | ···················· | ···················· | ····················· | ···················· | ·NN················· | ···················· | |
| L | 4849 | hCoV-19/Switzerland/BS-42395273/2020\|EPI_ISL_58195 | ···················· | ···················· | ····················· | ···················· | ·NNN················ | ···················· | 1 |
| L | 4851 | hCoV-19/Switzerland/BS-42396265/2020\|EPI_ISL_58195 | ···················· | ···················· | ····················· | ···················· | ···················· | ···················· | |
| L | 4852 | hCoV-19/Switzerland/BS-65242186/2020\|EPI_ISL_58195 | ···················· | ···················· | ····················· | ···················· | ···················· | ···················· | |
| L | 4856 | hCoV-19/Switzerland/BS-42397125/2020\|EPI_ISL_58196 | ···················· | ···················· | ····················· | ···················· | ·NN················· | ···················· | |
| L | 4857 | hCoV-19/Switzerland/BS-42400127/2020\|EPI_ISL_58197 | ···················· | ···················· | ····················· | ···················· | ·NN················· | ···················· | |
| L | 4859 | hCoV-19/Switzerland/BS-42404670/2020\|EPI_ISL_58197 | ···················· | ···················· | ····················· | ···················· | NNN················· | ···················· | 1 |
| L | 4865 | hCoV-19/USA/VA-DCLS-1459/2020\|EPI_ISL_572200\|2020- | ···················· | ···················· | ····················· | ···················· | ···················· | ···················· | |
| L | 4866 | hCoV-19/USA/VA-DCLS-1460/2020\|EPI_ISL_572201\|2020- | ···················· | ···················· | ····················· | ···················· | ···················· | ···················· | |
| L | 4867 | hCoV-19/USA/VA-DCLS-1461/2020\|EPI_ISL_572202\|2020- | ···················· | ···················· | ····················· | ···················· | ···················· | ···················· | |
| L | 4868 | hCoV-19/USA/VA-DCLS-1462/2020\|EPI_ISL_572203\|2020- | ···················· | ···················· | ····················· | ···················· | ···················· | ···················· | |
| L | 4869 | hCoV-19/USA/VA-DCLS-1463/2020\|EPI_ISL_572204\|2020- | ···················· | ···················· | ····················· | ···················· | ···················· | ···················· | |
| L | 4870 | hCoV-19/USA/VA-DCLS-1465/2020\|EPI_ISL_572205\|2020- | ···················· | ···················· | ····················· | ···················· | ···················· | ···················· | |
| L | 4871 | hCoV-19/USA/VA-DCLS-1466/2020\|EPI_ISL_572206\|2020- | ···················· | ···················· | ····················· | ···················· | ···················· | ···················· | |
| L | 4872 | hCoV-19/USA/VA-DCLS-1468/2020\|EPI_ISL_572207\|2020- | ···················· | ···················· | ····················· | ···················· | ···················· | ···················· | |
| L | 4873 | hCoV-19/USA/VA-DCLS-1469/2020\|EPI_ISL_572208\|2020- | ···················· | ···················· | ····················· | ···················· | ···················· | ···················· | |
| L | 4874 | hCoV-19/USA/VA-DCLS-1470/2020\|EPI_ISL_572209\|2020- | ···················· | ···················· | ····················· | ···················· | ···················· | ···················· | |
| L | 4875 | hCoV-19/USA/VA-DCLS-1471/2020\|EPI_ISL_572210\|2020- | ···················· | ···················· | ····················· | ···················· | ···················· | ···················· | |
| L | 4876 | hCoV-19/USA/VA-DCLS-1472/2020\|EPI_ISL_572211\|2020- | ···················· | ···················· | ····················· | ···················· | ···················· | ···················· | |
| L | 4877 | hCoV-19/USA/VA-DCLS-1473/2020\|EPI_ISL_572212\|2020- | ···················· | ···················· | ····················· | ···················· | ···················· | ···················· | |
| L | 4878 | hCoV-19/USA/VA-DCLS-1475/2020\|EPI_ISL_572213\|2020- | ···················· | ···················· | ····················· | ···················· | ···················· | ···················· | |
| L | 4879 | hCoV-19/USA/VA-DCLS-1476/2020\|EPI_ISL_572214\|2020- | ···················· | ···················· | ····················· | ···················· | ···················· | ···················· | |
| L | 4880 | hCoV-19/USA/VA-DCLS-1477/2020\|EPI_ISL_572215\|2020- | ···················· | ···················· | ····················· | ···················· | ···················· | ···················· | |
| L | 4881 | hCoV-19/USA/VA-DCLS-1478/2020\|EPI_ISL_572216\|2020- | ···················· | ···················· | ····················· | ···················· | ···················· | ···················· | |
| L | 4882 | hCoV-19/USA/VA-DCLS-1479/2020\|EPI_ISL_572217\|2020- | ···················· | ···················· | ····················· | ···················· | ···················· | ···················· | |
| L | 4883 | hCoV-19/USA/VA-DCLS-1677/2020\|EPI_ISL_572218\|2020- | ···················· | ···················· | ····················· | ···················· | ···················· | ···················· | |
| L | 4884 | hCoV-19/USA/VA-DCLS-1678/2020\|EPI_ISL_572219\|2020- | ···················· | ···················· | ····················· | ···················· | ···················· | ···················· | |
| L | 4885 | hCoV-19/USA/VA-DCLS-1376/2020\|EPI_ISL_572220\|2020- | ···················· | ···················· | ····················· | ···················· | ···················· | ···················· | |
| L | 4886 | hCoV-19/USA/VA-DCLS-1362/2020\|EPI_ISL_572221\|2020- | ···················· | ···················· | ····················· | ···················· | ···················· | ···················· | |
| L | 4887 | hCoV-19/USA/VA-DCLS-1363/2020\|EPI_ISL_572222\|2020- | ···················· | ···················· | ····················· | ···················· | ···················· | ···················· | |
| L | 4888 | hCoV-19/USA/VA-DCLS-1364/2020\|EPI_ISL_572223\|2020- | ···················· | ···················· | ····················· | ···················· | ···················· | ···················· | |
| L | 4889 | hCoV-19/USA/VA-DCLS-1371/2020\|EPI_ISL_572224\|2020- | ···················· | ···················· | ····················· | ···················· | ···················· | ···················· | |
| L | 4890 | hCoV-19/USA/VA-DCLS-1380/2020\|EPI_ISL_572225\|2020- | ···················· | ···················· | ····················· | ···················· | ···················· | ···················· | |
| L | 4891 | hCoV-19/USA/VA-DCLS-1381/2020\|EPI_ISL_572226\|2020- | ···················· | ···················· | ····················· | ···················· | ···················· | ···················· | |
| L | 4892 | hCoV-19/USA/VA-DCLS-1382/2020\|EPI_ISL_572227\|2020- | ···················· | ···················· | ····················· | ···················· | ···················· | ···················· | |
| L | 4893 | hCoV-19/USA/VA-DCLS-1383/2020\|EPI_ISL_572228\|2020- | ···················· | ···················· | ····················· | ···················· | ···················· | ···················· | |
| L | 4894 | hCoV-19/USA/VA-DCLS-1384/2020\|EPI_ISL_572229\|2020- | ···················· | ···················· | ····················· | ···················· | ···················· | ···················· | |
| L | 4895 | hCoV-19/USA/VA-DCLS-1387/2020\|EPI_ISL_572230\|2020- | ···················· | ·A·················· | ····················· | ···················· | ···················· | ···················· | |

| | | | | | | | | |
|---|---|---|---|---|---|---|---|---|
| L | 4896 | hCoV-19/USA/VA-DCLS-1388/2020\|EPI_ISL_572231\|2020- | ···················· | ···················· | ···················· | ···················· | ···················· | ········A··········· |
| L | 4897 | hCoV-19/USA/VA-DCLS-1390/2020\|EPI_ISL_572232\|2020- | ···················· | ···················· | ···················· | ···················· | ···················· | ···················· |
| L | 4898 | hCoV-19/USA/VA-DCLS-1391/2020\|EPI_ISL_572233\|2020- | ···················· | ···················· | ···················· | ···················· | ···················· | ···················· |
| L | 4899 | hCoV-19/USA/VA-DCLS-1392/2020\|EPI_ISL_572234\|2020- | ···················· | ···················· | ···················· | ···················· | ···················· | ···················· |
| L | 4900 | hCoV-19/USA/VA-DCLS-1394/2020\|EPI_ISL_572235\|2020- | ···················· | ···················· | ···················· | ···················· | ···················· | ···················· |
| L | 4901 | hCoV-19/USA/VA-DCLS-1395/2020\|EPI_ISL_572236\|2020- | ···················· | ·A·················· | ···················· | ···················· | ···················· | ···················· |
| L | 4902 | hCoV-19/USA/VA-DCLS-1396/2020\|EPI_ISL_572237\|2020- | ···················· | ·A·················· | ···················· | ···················· | ···················· | ···················· |
| L | 4903 | hCoV-19/USA/VA-DCLS-1398/2020\|EPI_ISL_572238\|2020- | ···················· | ···················· | ···················· | ···················· | ···················· | ···················· |
| L | 4904 | hCoV-19/USA/VA-DCLS-1400/2020\|EPI_ISL_572239\|2020- | ···················· | ···················· | ···················· | ···················· | ···················· | ···················· |
| L | 4905 | hCoV-19/USA/VA-DCLS-1402/2020\|EPI_ISL_572240\|2020- | ···················· | ···················· | ···················· | ···················· | ···················· | ···················· |
| L | 4906 | hCoV-19/USA/VA-DCLS-1404/2020\|EPI_ISL_572241\|2020- | ···················· | ···················· | ···················· | ···················· | ···················· | ···················· |
| L | 4907 | hCoV-19/USA/VA-DCLS-1405/2020\|EPI_ISL_572242\|2020- | ···················· | ···················· | ···················· | ···················· | ···················· | ········A··········· |
| L | 4908 | hCoV-19/USA/VA-DCLS-1406/2020\|EPI_ISL_572243\|2020- | ···················· | ···················· | ···················· | ···················· | ···················· | ···················· |
| L | 4909 | hCoV-19/USA/VA-DCLS-1409/2020\|EPI_ISL_572244\|2020- | ···················· | ···················· | ···················· | ···················· | ···················· | ···················· |
| L | 4910 | hCoV-19/USA/VA-DCLS-1410/2020\|EPI_ISL_572245\|2020- | ···················· | ···················· | ···················· | ···················· | ···················· | ···················· |
| L | 4911 | hCoV-19/USA/VA-DCLS-1308/2020\|EPI_ISL_572248\|2020- | ···················· | ···················· | ···················· | ···················· | ···················· | ···················· |
| L | 4912 | hCoV-19/USA/VA-DCLS-1324/2020\|EPI_ISL_572249\|2020- | ···················· | ···················· | ···················· | ···················· | ···················· | ···················· |
| L | 4913 | hCoV-19/USA/VA-DCLS-1315/2020\|EPI_ISL_572250\|2020- | ···················· | ···················· | ···················· | ···················· | ···················· | ···················· |
| L | 4914 | hCoV-19/USA/VA-DCLS-1323/2020\|EPI_ISL_572251\|2020- | ···················· | ···················· | ···················· | ···················· | ···················· | ···················· |
| L | 4915 | hCoV-19/USA/VA-DCLS-1310/2020\|EPI_ISL_572252\|2020- | ···················· | ···················· | ···················· | ···················· | ···················· | ···················· |
| L | 4916 | hCoV-19/USA/VA-DCLS-1311/2020\|EPI_ISL_572253\|2020- | ···················· | ···················· | ···················· | ···················· | ···················· | ···················· |
| L | 4917 | hCoV-19/USA/VA-DCLS-1312/2020\|EPI_ISL_572254\|2020- | ···················· | ···················· | ···················· | ···················· | ···················· | ···················· |
| L | 4918 | hCoV-19/USA/VA-DCLS-1313/2020\|EPI_ISL_572255\|2020- | ···················· | ···················· | ···················· | ···················· | ···················· | ···················· |
| L | 4919 | hCoV-19/USA/VA-DCLS-1314/2020\|EPI_ISL_572256\|2020- | ···················· | ···················· | ···················· | ···················· | ···················· | ···················· |
| L | 4920 | hCoV-19/USA/VA-DCLS-1317/2020\|EPI_ISL_572257\|2020- | ···················· | ···················· | ···················· | ···················· | ···················· | ···················· |
| L | 4921 | hCoV-19/USA/VA-DCLS-1327/2020\|EPI_ISL_572258\|2020- | ···················· | ···················· | ···················· | ···················· | ···················· | ···················· |
| L | 4922 | hCoV-19/USA/VA-DCLS-1328/2020\|EPI_ISL_572259\|2020- | ···················· | ···················· | ···················· | ···················· | ···················· | ···················· |
| L | 4923 | hCoV-19/USA/VA-DCLS-1331/2020\|EPI_ISL_572260\|2020- | ···················· | ···················· | ···················· | ···················· | ···················· | ···················· |
| L | 4924 | hCoV-19/USA/VA-DCLS-1332/2020\|EPI_ISL_572261\|2020- | ···················· | ···················· | ···················· | ···················· | ···················· | ···················· |
| L | 4925 | hCoV-19/USA/VA-DCLS-1337/2020\|EPI_ISL_572262\|2020- | ···················· | ···················· | ···················· | ···················· | ···················· | ···················· |
| L | 4926 | hCoV-19/USA/VA-DCLS-1339/2020\|EPI_ISL_572263\|2020- | ···················· | ···················· | ···················· | ···················· | ···················· | ···················· |
| L | 4927 | hCoV-19/USA/VA-DCLS-1343/2020\|EPI_ISL_572264\|2020- | ···················· | ···················· | ···················· | ···················· | ···················· | ···················· |
| L | 4928 | hCoV-19/USA/VA-DCLS-1346/2020\|EPI_ISL_572265\|2020- | ···················· | ···················· | ···················· | ···················· | ···················· | ···················· |
| L | 4929 | hCoV-19/England/QEUH-9AEE3E/2020\|EPI_ISL_573510\|20 | ···················· | ···················· | ···················· | ···················· | ···················· | ···················· |
| L | 4930 | hCoV-19/England/QEUH-9AEE4D/2020\|EPI_ISL_573511\|20 | ···················· | ···················· | ···················· | ···················· | ···················· | ···················· |
| L | 4931 | hCoV-19/England/QEUH-9AEE7A/2020\|EPI_ISL_573512\|20 | ···················· | ···················· | ···················· | ···················· | ···················· | ···················· |
| L | 4932 | hCoV-19/England/QEUH-9AEEF2/2020\|EPI_ISL_573513\|20 | ···················· | ···················· | ···················· | ···················· | ···T················ | ···················· |
| L | 4933 | hCoV-19/England/QEUH-9AEF2C/2020\|EPI_ISL_573514\|20 | ···················· | ···················· | ···················· | ···················· | ···T················ | ···················· |
| L | 4934 | hCoV-19/England/QEUH-9AEF3B/2020\|EPI_ISL_573515\|20 | ···················· | ···················· | ···················· | ···················· | ···T················ | ···················· |
| L | 4935 | hCoV-19/England/QEUH-9AEF4A/2020\|EPI_ISL_573516\|20 | ···················· | ···················· | ···················· | ···················· | ···T················ | ···················· |
| L | 4936 | hCoV-19/England/QEUH-9AEF68/2020\|EPI_ISL_573517\|20 | ···················· | ···················· | ···················· | ···················· | ···················· | ···················· |
| L | 4937 | hCoV-19/England/QEUH-9AEF77/2020\|EPI_ISL_573518\|20 | ···················· | ···················· | ···················· | ···················· | ···················· | ···················· |
| L | 4938 | hCoV-19/England/QEUH-9AEF86/2020\|EPI_ISL_573519\|20 | ···················· | ···················· | ···················· | ···················· | ···T················ | ···················· |
| L | 4939 | hCoV-19/England/QEUH-9AEF95/2020\|EPI_ISL_573520\|20 | ···················· | ···················· | ···················· | ···················· | ···················· | ···················· |
| L | 4940 | hCoV-19/England/QEUH-9AEFB3/2020\|EPI_ISL_573521\|20 | ···················· | ···················· | ···················· | ···················· | ···················· | ···················· |
| L | 4941 | hCoV-19/England/QEUH-9AEFC2/2020\|EPI_ISL_573522\|20 | ···················· | ···················· | ···················· | ···················· | ···T················ | ···················· |
| L | 4942 | hCoV-19/England/QEUH-9AEFD1/2020\|EPI_ISL_573523\|20 | ···················· | ···················· | ···················· | ···················· | ···················· | ···················· |
| L | 4943 | hCoV-19/England/QEUH-9AEFE0/2020\|EPI_ISL_573524\|20 | ···················· | ·A·················· | ···················· | ···················· | ···················· | ···················· |
| L | 4944 | hCoV-19/England/QEUH-9AEFFF/2020\|EPI_ISL_573525\|20 | ···················· | ···················· | ···················· | ···················· | ···T················ | ···················· |
| L | 4945 | hCoV-19/England/QEUH-9B3A7E/2020\|EPI_ISL_573526\|20 | ···················· | ···················· | ···················· | ···················· | ···················· | ···················· |
| L | 4946 | hCoV-19/England/QEUH-9B3ABA/2020\|EPI_ISL_573527\|20 | ···················· | ···················· | ···················· | ···················· | ···················· | ···················· |
| L | 4947 | hCoV-19/England/QEUH-9B3AD8/2020\|EPI_ISL_573528\|20 | ···················· | ···················· | ···················· | ···················· | ···················· | ···················· |
| L | 4948 | hCoV-19/England/QEUH-9B3AF6/2020\|EPI_ISL_573529\|20 | ···················· | ···················· | ···················· | ···················· | ···················· | ···················· |
| L | 4949 | hCoV-19/England/QEUH-9B3B02/2020\|EPI_ISL_573530\|20 | ···················· | ···················· | ···················· | ···················· | ···················· | ···················· |
| L | 4950 | hCoV-19/England/QEUH-9B3B11/2020\|EPI_ISL_573531\|20 | ···················· | ···················· | ········T··········· | ···················· | ·····T·············· | ···················· |
| L | 4951 | hCoV-19/England/QEUH-9B3B20/2020\|EPI_ISL_573532\|20 | ···················· | ···················· | ···················· | ···················· | ···················· | ···················· |
| L | 4952 | hCoV-19/England/QEUH-9B3B3F/2020\|EPI_ISL_573533\|20 | ···················· | ···················· | ···················· | ···················· | ···················· | ···················· |
| L | 4953 | hCoV-19/England/QEUH-9B3B4E/2020\|EPI_ISL_573534\|20 | ···················· | ···················· | ···················· | ···················· | ···················· | ···················· |
| L | 4954 | hCoV-19/England/QEUH-9B3B8A/2020\|EPI_ISL_573535\|20 | ···················· | ···················· | ···················· | ···················· | ···T················ | ···················· |
| L | 4955 | hCoV-19/England/QEUH-9B3B99/2020\|EPI_ISL_573536\|20 | ···················· | ···················· | ···················· | ···················· | ···················· | ···················· |
| L | 4956 | hCoV-19/England/QEUH-9B3BA8/2020\|EPI_ISL_573537\|20 | ···················· | ···················· | ···················· | ···················· | ···················· | ···················· |
| L | 4957 | hCoV-19/England/QEUH-9B3BC6/2020\|EPI_ISL_573538\|20 | ···················· | ···················· | ···················· | ···················· | ···················· | ···················· |
| L | 4958 | hCoV-19/England/QEUH-9B3BE4/2020\|EPI_ISL_573539\|20 | ···················· | ···················· | ···················· | ···················· | ···················· | ···················· |
| L | 4959 | hCoV-19/England/QEUH-9B3C0F/2020\|EPI_ISL_573540\|20 | ···················· | ·A·················· | ···················· | ···················· | ···················· | ··A················· |
| L | 4960 | hCoV-19/England/QEUH-9B3C1E/2020\|EPI_ISL_573541\|20 | ···················· | ···················· | ···················· | ···················· | ···················· | ···················· |
| L | 4961 | hCoV-19/England/QEUH-9B3C2D/2020\|EPI_ISL_573542\|20 | ···················· | ···················· | ···················· | ···················· | ···················· | ···················· |
| L | 4962 | hCoV-19/England/QEUH-9B3C4B/2020\|EPI_ISL_573543\|20 | ···················· | ···················· | ···················· | ···················· | ···················· | ···················· |
| L | 4963 | hCoV-19/England/QEUH-9B3CB4/2020\|EPI_ISL_573544\|20 | ···················· | ···················· | ···················· | ···················· | ···················· | ···················· |
| L | 4964 | hCoV-19/England/QEUH-9B3CF0/2020\|EPI_ISL_573545\|20 | ···················· | ···················· | ···················· | ···················· | ···················· | ···················· |
| L | 4965 | hCoV-19/England/QEUH-9B3D2A/2020\|EPI_ISL_573546\|20 | ···················· | ···················· | ···················· | ···················· | ···················· | ···················· |
| L | 4966 | hCoV-19/England/QEUH-9B3D48/2020\|EPI_ISL_573547\|20 | ···················· | ···················· | ···················· | ···················· | ···················· | ···················· |
| L | 4967 | hCoV-19/England/QEUH-9B3D57/2020\|EPI_ISL_573548\|20 | ···················· | ···················· | ···················· | ···················· | ···················· | ···················· |
| L | 4968 | hCoV-19/England/QEUH-9B3D84/2020\|EPI_ISL_573549\|20 | ···················· | ···················· | ···················· | ···················· | ···················· | ···················· |
| L | 4969 | hCoV-19/England/QEUH-9B3DEE/2020\|EPI_ISL_573550\|20 | ···················· | ···················· | ···················· | ···················· | ···················· | ···················· |
| L | 4970 | hCoV-19/England/QEUH-9B3E18/2020\|EPI_ISL_573551\|20 | ···················· | ···················· | ···················· | ···················· | ···················· | ···················· |
| L | 4971 | hCoV-19/England/QEUH-9B3E45/2020\|EPI_ISL_573552\|20 | ···················· | ···················· | ···················· | ···················· | ···················· | ···················· |
| L | 4972 | hCoV-19/England/QEUH-9B3E63/2020\|EPI_ISL_573553\|20 | ···················· | ·A·················· | ···················· | ···················· | ···················· | ···················· |
| L | 4973 | hCoV-19/England/QEUH-9B3E72/2020\|EPI_ISL_573554\|20 | ···················· | ···················· | ···················· | ···················· | ···················· | ···················· |
| L | 4974 | hCoV-19/England/QEUH-9B3E90/2020\|EPI_ISL_573555\|20 | ···················· | ·A·················· | ···················· | ···················· | ···················· | ···················· |
| L | 4975 | hCoV-19/England/QEUH-9B3EAF/2020\|EPI_ISL_573556\|20 | ···················· | ···················· | ···················· | ···················· | ···················· | ···················· |

| | | | | | | | | |
|---|---|---|---|---|---|---|---|---|
| L | 4976 | hCoV-19/England/QEUH-9B3EDC/2020\|EPI_ISL_573557\|20 | ···················· | ···················· | ····················· | ···················· | ···················· | ···················· |
| L | 4977 | hCoV-19/England/QEUH-9B3F06/2020\|EPI_ISL_573558\|20 | ···················· | ···················· | ····················· | ···················· | ···················· | ···················· |
| L | 4978 | hCoV-19/England/QEUH-9B3F33/2020\|EPI_ISL_573559\|20 | ···················· | ···················· | ····················· | ···················· | ···················· | ···················· |
| L | 4979 | hCoV-19/England/QEUH-9B3F8E/2020\|EPI_ISL_573560\|20 | ···················· | ···················· | ····················· | ···················· | ···················· | ···················· |
| L | 4980 | hCoV-19/England/QEUH-9B3F9D/2020\|EPI_ISL_573561\|20 | ···················· | ···················· | ····················· | ···················· | ···················· | ···················· |
| L | 4981 | hCoV-19/England/QEUH-9B3FE8/2020\|EPI_ISL_573562\|20 | ···················· | ·A·················· | ····················· | ···················· | ···················· | ··A················· |
| L | 4982 | hCoV-19/England/QEUH-9B4002/2020\|EPI_ISL_573563\|20 | ···················· | ···················· | ····················· | ···················· | ···················· | ···················· |
| L | 4983 | hCoV-19/England/QEUH-9B44D9/2020\|EPI_ISL_573564\|20 | ···················· | ···················· | ····················· | ···················· | ···················· | ···················· |
| L | 4984 | hCoV-19/England/QEUH-9B44E8/2020\|EPI_ISL_573565\|20 | ···················· | ·A·················· | ····················· | ···················· | ···················· | ··A················· |
| L | 4985 | hCoV-19/England/QEUH-9B9F4C/2020\|EPI_ISL_573566\|20 | ···················· | ···················· | ····················· | ···················· | ···················· | ···················· |
| L | 4986 | hCoV-19/England/QEUH-9B9F5B/2020\|EPI_ISL_573567\|20 | ···················· | ···················· | ····················· | ···················· | ···················· | ···················· |
| L | 4987 | hCoV-19/England/QEUH-9B9FC4/2020\|EPI_ISL_573568\|20 | ···················· | ···················· | ····················· | ···················· | ···················· | ···················· |
| L | 4988 | hCoV-19/England/QEUH-9B9FD3/2020\|EPI_ISL_573569\|20 | ···················· | ···················· | ····················· | ···················· | ···················· | ···················· |
| L | 4989 | hCoV-19/England/QEUH-9BA01B/2020\|EPI_ISL_573570\|20 | ···················· | ···················· | ····················· | ···················· | ···················· | ···················· |
| L | 4990 | hCoV-19/England/QEUH-9BA02A/2020\|EPI_ISL_573571\|20 | ···················· | ···················· | ····················· | ···················· | ···················· | ···················· |
| L | 4991 | hCoV-19/England/QEUH-9BA048/2020\|EPI_ISL_573572\|20 | ···················· | ···················· | ····················· | ···················· | ···················· | ···················· |
| L | 4992 | hCoV-19/England/QEUH-9BA066/2020\|EPI_ISL_573573\|20 | ···················· | ···················· | ····················· | ···················· | ···················· | ···················· |
| L | 4993 | hCoV-19/England/QEUH-9BA075/2020\|EPI_ISL_573574\|20 | ···················· | ···················· | ····················· | ···················· | ···················· | ···················· |
| L | 4994 | hCoV-19/England/QEUH-9BA084/2020\|EPI_ISL_573575\|20 | ···················· | ···················· | ····················· | ···················· | ···················· | ···················· |
| L | 4995 | hCoV-19/England/QEUH-9BA093/2020\|EPI_ISL_573576\|20 | ···················· | ···················· | ····················· | ···················· | ···················· | ···················· |
| L | 4996 | hCoV-19/England/QEUH-9BA0B1/2020\|EPI_ISL_573577\|20 | ···················· | ···················· | ····················· | ···················· | ···················· | ···················· |
| L | 4997 | hCoV-19/England/QEUH-9BA0EE/2020\|EPI_ISL_573578\|20 | ···················· | ···················· | ····················· | ···················· | ···················· | ···················· |
| L | 4998 | hCoV-19/England/QEUH-9BA109/2020\|EPI_ISL_573579\|20 | ···················· | ···················· | ····················· | ···················· | ···················· | ···················· |
| L | 4999 | hCoV-19/England/QEUH-9BA127/2020\|EPI_ISL_573580\|20 | ···················· | ···················· | ····················· | ···················· | ···················· | ···················· |
| L | 5000 | hCoV-19/England/QEUH-9BA136/2020\|EPI_ISL_573581\|20 | ···················· | ···················· | ····················· | ···················· | ···················· | ···················· |
| L | 5001 | hCoV-19/England/QEUH-9BA163/2020\|EPI_ISL_573582\|20 | ···················· | ···················· | ····················· | ···················· | ···················· | ···················· |
| L | 5002 | hCoV-19/England/QEUH-9BA172/2020\|EPI_ISL_573583\|20 | ···················· | ···················· | ····················· | ···················· | ···················· | ···················· |
| L | 5003 | hCoV-19/England/QEUH-9BA1AF/2020\|EPI_ISL_573584\|20 | ···················· | ···················· | ····················· | ···················· | ···················· | ···················· |
| L | 5004 | hCoV-19/England/QEUH-9BA1EB/2020\|EPI_ISL_573585\|20 | ···················· | ···················· | ····················· | ···················· | ···················· | ···················· |
| L | 5005 | hCoV-19/England/QEUH-9BA242/2020\|EPI_ISL_573586\|20 | ···················· | ···················· | ····················· | ···················· | ···················· | ···················· |
| L | 5006 | hCoV-19/England/QEUH-9BA2F7/2020\|EPI_ISL_573587\|20 | ···················· | ···················· | ····················· | ···················· | ···················· | ···················· |
| L | 5007 | hCoV-19/England/QEUH-9BA37C/2020\|EPI_ISL_573588\|20 | ···················· | ···················· | ····················· | ···················· | ···················· | ···················· |
| L | 5008 | hCoV-19/England/QEUH-9BA3D6/2020\|EPI_ISL_573589\|20 | ···················· | ···················· | ····················· | ···················· | ···················· | ···················· |
| L | 5009 | hCoV-19/England/QEUH-9BA479/2020\|EPI_ISL_573590\|20 | ···················· | ···················· | ····················· | ···················· | ···················· | ···················· |
| L | 5010 | hCoV-19/England/QEUH-9BA497/2020\|EPI_ISL_573591\|20 | ···················· | ·A·················· | ····················· | ···················· | ···················· | ··A················· |
| L | 5011 | hCoV-19/England/QEUH-9BA4A6/2020\|EPI_ISL_573592\|20 | ···················· | ···················· | ····················· | ···················· | ···················· | ···················· |
| L | 5012 | hCoV-19/England/QEUH-9BA4C4/2020\|EPI_ISL_573593\|20 | ···················· | ···················· | ····················· | ···················· | ···················· | ···················· |
| L | 5013 | hCoV-19/England/QEUH-9BA576/2020\|EPI_ISL_573594\|20 | ···················· | ···················· | ····················· | ···················· | ···················· | ···················· |
| L | 5014 | hCoV-19/England/QEUH-9BA5FE/2020\|EPI_ISL_573595\|20 | ···················· | ···················· | ····················· | ···················· | ···················· | ···················· |
| L | 5015 | hCoV-19/England/QEUH-9BA619/2020\|EPI_ISL_573596\|20 | ···················· | ···················· | ····················· | ···················· | ···················· | ···················· |
| L | 5016 | hCoV-19/Switzerland/BE-UNIBE-202-6410/2020\|EPI_ISL | ···················· | ···················· | ····················· | ···················· | ···················· | ···················· |
| L | 5017 | hCoV-19/Switzerland/BE-UNIBE-203-6441/2020\|EPI_ISL | ···················· | ···················· | ····················· | ···················· | ···················· | ···················· |
| L | 5018 | hCoV-19/Switzerland/BE-UNIBE-204-6477/2020\|EPI_ISL | ···················· | ·A·················· | ····················· | ···················· | ···················· | ···················· |
| L | 5019 | hCoV-19/Switzerland/BE-UNIBE-205-7690/2020\|EPI_ISL | ···················· | ···················· | ····················· | ···················· | ···················· | ···················· |
| L | 5020 | hCoV-19/Switzerland/BE-UNIBE-206-7951/2020\|EPI_ISL | ···················· | ···················· | ····················· | ···················· | ···················· | ···················· |
| L | 5021 | hCoV-19/Switzerland/BE-UNIBE-207-8159/2020\|EPI_ISL | ···················· | ···················· | ····················· | ···················· | ···················· | ···················· |
| L | 5022 | hCoV-19/Switzerland/BE-UNIBE-208-8169/2020\|EPI_ISL | ···················· | ···················· | ····················· | ···················· | ···················· | ···················· |
| L | 5023 | hCoV-19/Switzerland/BE-UNIBE-209-8650/2020\|EPI_ISL | ···················· | ···················· | ····················· | ···················· | ···················· | ···················· |
| L | 5024 | hCoV-19/Switzerland/BE-UNIBE-210-8666/2020\|EPI_ISL | ···················· | ···················· | ····················· | ···················· | ···················· | ···················· |
| L | 5025 | hCoV-19/Switzerland/BE-UNIBE-211-9026/2020\|EPI_ISL | ···················· | ···················· | ····················· | ···················· | ···················· | ···················· |
| L | 5026 | hCoV-19/Switzerland/BE-UNIBE-212-9055/2020\|EPI_ISL | ···················· | ·A·················· | ····················· | ···················· | ···················· | ···················· |
| L | 5027 | hCoV-19/Switzerland/BE-UNIBE-214-0044/2020\|EPI_ISL | ···················· | ···················· | ····················· | ···················· | ···················· | ···················· |
| L | 5028 | hCoV-19/Switzerland/BE-UNIBE-215-0080/2020\|EPI_ISL | ···················· | ·A·················· | ····················· | ···················· | ···················· | ···················· |
| L | 5029 | hCoV-19/Switzerland/BE-UNIBE-217-0518/2020\|EPI_ISL | ···················· | ···················· | ····················· | ···················· | ···················· | ···················· |
| L | 5030 | hCoV-19/Switzerland/BE-UNIBE-218-0537/2020\|EPI_ISL | ···················· | ···················· | ····················· | ···················· | ···················· | ···················· |
| L | 5031 | hCoV-19/Switzerland/BE-UNIBE-219-1251/2020\|EPI_ISL | ···················· | ···················· | ····················· | ···················· | ···················· | ···················· |
| L | 5032 | hCoV-19/Switzerland/BE-UNIBE-220-1253/2020\|EPI_ISL | ···················· | ···················· | ····················· | ···················· | ···················· | ···················· |
| L | 5033 | hCoV-19/Switzerland/BE-UNIBE-222-1255/2020\|EPI_ISL | ···················· | ···················· | ····················· | ···················· | ···················· | ···················· |
| L | 5034 | hCoV-19/Switzerland/BE-UNIBE-223-2673/2020\|EPI_ISL | ···················· | ···················· | ····················· | ···················· | ···················· | ···················· |
| L | 5035 | hCoV-19/Switzerland/BE-UNIBE-225-2890/2020\|EPI_ISL | ···················· | ···················· | ····················· | ···················· | ···················· | ···················· |
| L | 5036 | hCoV-19/Switzerland/BE-UNIBE-226-3606/2020\|EPI_ISL | ···················· | ···················· | ····················· | ···················· | ···················· | ···················· |
| L | 5037 | hCoV-19/Switzerland/BE-UNIBE-227-3922/2020\|EPI_ISL | ···················· | ···················· | ····················· | ···················· | ···················· | ···················· |
| L | 5038 | hCoV-19/Switzerland/BE-UNIBE-228-9106/2020\|EPI_ISL | ···················· | ···················· | ····················· | ···················· | ···················· | ···················· |
| L | 5039 | hCoV-19/Switzerland/AG-ETHZ-100193/2020\|EPI_ISL_57 | ···················· | ···················· | ····················· | ···················· | ···················· | ···················· |
| L | 5040 | hCoV-19/Switzerland/AG-ETHZ-100217/2020\|EPI_ISL_57 | ···················· | ···················· | ····················· | ···················· | ···················· | ···················· |
| L | 5041 | hCoV-19/Switzerland/AG-ETHZ-100224/2020\|EPI_ISL_57 | ···················· | ···················· | ····················· | ···················· | ···················· | ···················· |
| L | 5042 | hCoV-19/Switzerland/AG-ETHZ-100306/2020\|EPI_ISL_57 | ···················· | ···················· | ····················· | ···················· | ···················· | ···················· |
| L | 5043 | hCoV-19/Switzerland/AG-ETHZ-100351/2020\|EPI_ISL_57 | ···················· | ···················· | ····················· | ···················· | ···················· | ···················· |
| L | 5044 | hCoV-19/Switzerland/AG-ETHZ-100379/2020\|EPI_ISL_57 | ···················· | ···················· | ····················· | ···················· | ···················· | ···················· |
| L | 5045 | hCoV-19/Switzerland/AG-ETHZ-100381/2020\|EPI_ISL_57 | ···················· | ···················· | ····················· | ···················· | ···················· | ···················· |
| L | 5046 | hCoV-19/Switzerland/BL-ETHZ-100210/2020\|EPI_ISL_57 | ···················· | ···················· | ····················· | ···················· | ·······S············ | ············R······· |
| L | 5047 | hCoV-19/Switzerland/BL-ETHZ-100234/2020\|EPI_ISL_57 | ···················· | ···················· | ····················· | ···················· | ···················· | ···················· |
| L | 5048 | hCoV-19/Switzerland/BL-ETHZ-100235/2020\|EPI_ISL_57 | ···················· | ···················· | ····················· | ···················· | ···················· | ···················· |
| L | 5049 | hCoV-19/Switzerland/BL-ETHZ-100236/2020\|EPI_ISL_57 | ···················· | ···················· | ····················· | ···················· | ···················· | ···················· |
| L | 5050 | hCoV-19/Switzerland/BL-ETHZ-100244/2020\|EPI_ISL_57 | ···················· | ···················· | ····················· | ···················· | ···················· | ···················· |
| L | 5051 | hCoV-19/Switzerland/BL-ETHZ-100304/2020\|EPI_ISL_57 | ···················· | ···················· | ····················· | ···················· | ···················· | ···················· |
| L | 5052 | hCoV-19/Switzerland/BL-ETHZ-100305/2020\|EPI_ISL_57 | ···················· | ·A·················· | ····················· | ···················· | ···················· | ···················· |
| L | 5053 | hCoV-19/Switzerland/BL-ETHZ-100308/2020\|EPI_ISL_57 | ···················· | ···················· | ····················· | ···················· | ···················· | ···················· |
| L | 5054 | hCoV-19/Switzerland/BL-ETHZ-100311/2020\|EPI_ISL_57 | ···················· | ···················· | ····················· | ···················· | ···················· | ···················· |
| L | 5055 | hCoV-19/Switzerland/BL-ETHZ-100342/2020\|EPI_ISL_57 | ···················· | ···················· | ····················· | ···················· | ···················· | ···················· |

L 5056 hCoV-19/Switzerland/BL-ETHZ-100343/2020|EPI_ISL_57 ···················· ···················· ···················· ···················· ···················· ····················
L 5057 hCoV-19/Switzerland/BL-ETHZ-100344/2020|EPI_ISL_57 ···················· ···················· ···················· ···················· ···················· ····················
L 5058 hCoV-19/Switzerland/BL-ETHZ-100345/2020|EPI_ISL_57 ···················· ···················· ···················· ···················· ···················· ····················
L 5059 hCoV-19/Switzerland/BL-ETHZ-100346/2020|EPI_ISL_57 ···················· ···················· ···················· ···················· ···················· ····················
L 5060 hCoV-19/Switzerland/BL-ETHZ-100347/2020|EPI_ISL_57 ···················· ···················· ···················· ···················· ···················· ····················
L 5061 hCoV-19/Switzerland/BL-ETHZ-100350/2020|EPI_ISL_57 ···················· ···················· ···················· ···················· ···················· ····················
L 5062 hCoV-19/Ireland/CO-NVRL-73IRL40054/2020|EPI_ISL_57 ···················· ···················· ···················· ···················· ···················· ····················
L 5063 hCoV-19/Ireland/D-NVRL-20ILR05740/2020|EPI_ISL_578 ···················· ·A·················· ···················· ···················· ···················· ···A················
L 5065 hCoV-19/Ireland/D-NVRL-72IRL36222/2020|EPI_ISL_578 ···················· ···················· ···················· ···················· ···················· ····················
L 5066 hCoV-19/Ireland/D-NVRL-72IRL41325/2020|EPI_ISL_578 ···················· ···················· ···················· ···················· ···················· ····················
L 5067 hCoV-19/Ireland/D-NVRL-73IRL14028/2020|EPI_ISL_578 ···················· ···················· ···················· ···················· ···················· ····················
L 5068 hCoV-19/Ireland/D-NVRL-73IRL22417/2020|EPI_ISL_578 ···················· ···················· ···················· ···················· ···················· ····················
L 5069 hCoV-19/Ireland/D-NVRL-73IRL22431/2020|EPI_ISL_578 ···················· ···················· ···················· ···················· ···················· ····················
L 5070 hCoV-19/Ireland/D-NVRL-73IRL25053/2020|EPI_ISL_578 ···················· ···················· ···················· ···················· ···················· ····················
L 5071 hCoV-19/Ireland/D-NVRL-73IRL25093/2020|EPI_ISL_578 ···················· ···················· ···················· ···················· ···················· ····················
L 5072 hCoV-19/Ireland/D-NVRL-73IRL25386/2020|EPI_ISL_578 ···················· ···················· ···················· ···················· ···················· ····················
L 5073 hCoV-19/Ireland/D-NVRL-73IRL25596/2020|EPI_ISL_578 ···················· ···················· ···················· ···················· ···················· ····················
L 5074 hCoV-19/Ireland/D-NVRL-73IRL26673/2020|EPI_ISL_578 ···················· ···················· ···················· ···················· ···················· ····················
L 5075 hCoV-19/Ireland/D-NVRL-73IRL27308/2020|EPI_ISL_578 ···················· ···················· ···················· ···················· ···················· ····················
L 5076 hCoV-19/Ireland/D-NVRL-73IRL27703/2020|EPI_ISL_578 ···················· ···················· ···················· ···················· ···················· ····················
L 5077 hCoV-19/Ireland/D-NVRL-73IRL27709/2020|EPI_ISL_578 ···················· ···················· ···················· ···················· ···················· ····················
L 5079 hCoV-19/USA/WI-WSLH-200175/2020|EPI_ISL_578401|202 ···················· ···················· ···················· ···················· ···················· ····················
L 5082 hCoV-19/Canada/NS-NML-955/2020|EPI_ISL_579585|2020 ···················· ···················· ···················· ···················· ···················· ····················
L 5083 hCoV-19/Canada/NS-NML-960/2020|EPI_ISL_579587|2020 ···················· ···················· ···················· ···················· ···················· ····················
L 5084 hCoV-19/Canada/NS-NML-961/2020|EPI_ISL_579588|2020 ···················· ···················· ···················· ···················· ···················· ····················
L 5085 hCoV-19/Canada/NS-NML-963/2020|EPI_ISL_579589|2020 ···················· ···················· ···················· ···················· ···················· ····················
L 5086 hCoV-19/Canada/NS-NML-964/2020|EPI_ISL_579590|2020 ···················· ···················· ···················· ···················· ···················· ····················
L 5087 hCoV-19/Canada/NS-NML-965/2020|EPI_ISL_579591|2020 ···················· ···················· ···················· ···················· ···················· ····················
L 5088 hCoV-19/Canada/NS-NML-966/2020|EPI_ISL_579592|2020 ···················· ···················· ···················· ···················· ···················· ····················
L 5089 hCoV-19/Canada/NS-NML-968/2020|EPI_ISL_579593|2020 ···················· ···················· ···················· ···················· ···················· ····················
L 5090 hCoV-19/Canada/NS-NML-1196/2020|EPI_ISL_579597|202 ···················· ···················· ···················· ···················· ···················· ····················
L 5091 hCoV-19/Canada/NS-NML-1197/2020|EPI_ISL_579598|202 ···················· ···················· ···················· ···················· ···················· ····················
L 5092 hCoV-19/England/CAMC-9EC10E/2020|EPI_ISL_580582|20 ···················· ···················· ···················· ···················· ···················· ····················
L 5093 hCoV-19/England/ALDP-9E66AB/2020|EPI_ISL_580583|20 ···················· ···················· ···················· ···················· ···················· ····················
L 5094 hCoV-19/England/ALDP-9E7103/2020|EPI_ISL_580584|20 ···················· ···················· ···················· ···················· ···················· ····················
L 5095 hCoV-19/England/ALDP-9E70BB/2020|EPI_ISL_580585|20 ···················· ···················· ···················· ···················· ···················· ····················
L 5096 hCoV-19/England/CAMC-9EC757/2020|EPI_ISL_580586|20 ···················· ···················· ···················· ···················· ···················· ····················
L 5097 hCoV-19/England/CAMC-9EC344/2020|EPI_ISL_580587|20 ···················· ···················· ···················· ···················· ···················· ····················
L 5098 hCoV-19/England/ALDP-9E6289/2020|EPI_ISL_580588|20 ···················· ···················· ···················· ···················· ···················· ····················
L 5099 hCoV-19/England/ALDP-9E730D/2020|EPI_ISL_580589|20 ···················· ···················· ···················· ···················· ···················· ····················
L 5100 hCoV-19/England/ALDP-9E625C/2020|EPI_ISL_580591|20 ···················· ···················· ···················· ···················· ···················· ····················
L 5101 hCoV-19/England/ALDP-9E6395/2020|EPI_ISL_580592|20 ···················· ···················· ···················· ···················· ···················· ····················
L 5102 hCoV-19/England/CAMC-9EC2ED/2020|EPI_ISL_580593|20 ···················· ···················· ···················· ···················· ···················· ····················
L 5103 hCoV-19/England/ALDP-9E6E58/2020|EPI_ISL_580595|20 ···················· ···················· ···················· ···················· ···················· ····················
L 5104 hCoV-19/England/ALDP-9E73C1/2020|EPI_ISL_580596|20 ···················· ···················· ···················· ···················· ···················· ····················
L 5105 hCoV-19/England/ALDP-9E6BD9/2020|EPI_ISL_580597|20 ···················· ···················· ···················· ···················· ···················· ····················
L 5106 hCoV-19/Wales/ALDP-9E6368/2020|EPI_ISL_580598|2020 ···················· ···················· ···················· ···················· ···················· ····················
L 5107 hCoV-19/England/ALDP-9E68C3/2020|EPI_ISL_580599|20 ···················· ···················· ···················· ···················· ···················· ····················
L 5108 hCoV-19/Wales/MILK-9F0662/2020|EPI_ISL_580700|2020 ···················· ···················· ···················· ···················· ···················· ····················
L 5109 hCoV-19/England/MILK-9EE7CE/2020|EPI_ISL_580701|20 ···················· ···················· ···················· ···················· ···················· ····················
L 5110 hCoV-19/England/MILK-9F043B/2020|EPI_ISL_580702|20 ···················· ···················· ···················· ···················· ···················· ····················
L 5111 hCoV-19/England/MILK-9F05ED/2020|EPI_ISL_580704|20 ···················· ···················· ···················· ···················· ···················· ····················
L 5112 hCoV-19/England/MILK-9EEC0B/2020|EPI_ISL_580709|20 ···················· ···················· ···················· ···················· ···················· ····················
L 5113 hCoV-19/England/ALDP-9EDDFA/2020|EPI_ISL_580710|20 ···················· ···················· ···················· ···················· ···················· ····················
L 5114 hCoV-19/Wales/MILK-9F06BD/2020|EPI_ISL_580711|2020 ···················· ···················· ···················· ···················· ···················· ····················
L 5115 hCoV-19/Wales/MILK-9F0635/2020|EPI_ISL_580712|2020 ···················· ···················· ···················· ···················· ···················· ····················
L 5116 hCoV-19/England/MILK-9F0574/2020|EPI_ISL_580713|20 ···················· ·A·······A·········· ···················· ···················· ···················· ····················
L 5117 hCoV-19/England/MILK-9EE403/2020|EPI_ISL_580716|20 ···················· ···················· ···················· ···················· ···················· ····················
L 5118 hCoV-19/England/MILK-9F0310/2020|EPI_ISL_580717|20 ···················· ···················· ···················· ···················· ···················· ····················
L 5119 hCoV-19/Wales/MILK-9F01CB/2020|EPI_ISL_580718|2020 ···················· ···················· ···················· ···················· ···················· ····················
L 5120 hCoV-19/Wales/MILK-9EEED8/2020|EPI_ISL_580719|2020 ···················· ···················· ···················· ···················· ···················· ····················
L 5121 hCoV-19/USA/MN-QDX-1361/2020|EPI_ISL_572150|2020-0 ···················· ···················· ···················· ···················· ···················· ····················
L 5122 hCoV-19/USA/MN-QDX-1362/2020|EPI_ISL_572151|2020-0 ···················· ···················· ···················· ···················· ···················· ····················
L 5123 hCoV-19/USA/MN-QDX-1363/2020|EPI_ISL_572152|2020-0 ···················· ···················· ···················· ···················· ···················· ····················
L 5124 hCoV-19/USA/MN-QDX-1364/2020|EPI_ISL_572153|2020-0 ···················· ···················· ···················· ···················· ···················· ····················
L 5125 hCoV-19/USA/MN-QDX-1365/2020|EPI_ISL_572154|2020-0 ···················· ···················· ···················· ···················· ···················· ····················
L 5126 hCoV-19/USA/MN-QDX-1366/2020|EPI_ISL_572155|2020-0 ···················· ···················· ···················· ···················· ···················· ····················
L 5127 hCoV-19/USA/MN-QDX-1367/2020|EPI_ISL_572156|2020-0 ···················· ···················· ···················· ···················· ···················· ····················
L 5128 hCoV-19/USA/MN-QDX-1368/2020|EPI_ISL_572157|2020-0 ···················· ···················· ···················· ···················· ···················· ····················
L 5129 hCoV-19/USA/MN-QDX-1369/2020|EPI_ISL_572158|2020-0 ···················· ···················· ···················· ···················· ···················· ····················
L 5130 hCoV-19/USA/MN-QDX-1370/2020|EPI_ISL_572159|2020-0 ···················· ···················· ···················· ···················· ···················· ····················
L 5131 hCoV-19/USA/MN-QDX-1371/2020|EPI_ISL_572160|2020-0 ···················· ···················· ···················· ···················· ···················· ····················
L 5132 hCoV-19/USA/MN-QDX-1372/2020|EPI_ISL_572161|2020-0 ···················· ···················· ···················· ···················· ···················· ····················
L 5133 hCoV-19/USA/MN-QDX-1373/2020|EPI_ISL_572162|2020-0 ···················· ···················· ···················· ···················· ···················· ····················
L 5134 hCoV-19/USA/MN-QDX-1374/2020|EPI_ISL_572163|2020-0 ···················· ···················· ···················· ···················· ···················· ····················
L 5135 hCoV-19/USA/MN-QDX-1375/2020|EPI_ISL_572164|2020-0 ···················· ···················· ···················· ···················· ···················· ····················
L 5136 hCoV-19/USA/MN-QDX-1376/2020|EPI_ISL_572165|2020-0 ···················· ···················· ···················· ···················· ···················· ····················
L 5137 hCoV-19/USA/MN-QDX-1377/2020|EPI_ISL_572166|2020-0 ···················· ···················· ···················· ···················· ···················· ····················
L 5138 hCoV-19/England/QEUH-9AEA76/2020|EPI_ISL_573478|20 ···················· ·A·················· ···················· ···················· ···················· ····················
L 5139 hCoV-19/England/QEUH-9AEB37/2020|EPI_ISL_573480|20 ···················· ···················· ···················· ···················· ···················· ····················

| | | | | | | | | |
|---|---|---|---|---|---|---|---|---|
| L | 5140 | hCoV-19/England/QEUH-9AEB46/2020\|EPI_ISL_573481\|20 | ···················· | ···················· | ····················· | ···················· | ···················· | ···················· |
| L | 5141 | hCoV-19/England/QEUH-9AEB64/2020\|EPI_ISL_573482\|20 | ···················· | ···················· | ····················· | ···················· | ···················· | ···················· |
| L | 5142 | hCoV-19/England/QEUH-9AEB73/2020\|EPI_ISL_573483\|20 | ···················· | ···················· | ····················· | ···················· | ···················· | ···················· |
| L | 5143 | hCoV-19/England/QEUH-9AEBBF/2020\|EPI_ISL_573484\|20 | ···················· | ·A·················· | ····················· | ···················· | ···················· | ···················· |
| L | 5144 | hCoV-19/England/QEUH-9AEBDD/2020\|EPI_ISL_573485\|20 | ···················· | ···················· | ····················· | ···················· | ···················· | ···················· |
| L | 5145 | hCoV-19/England/QEUH-9AEBEC/2020\|EPI_ISL_573486\|20 | ···················· | ···················· | ····················· | ···················· | ···················· | ···················· |
| L | 5146 | hCoV-19/England/QEUH-9AEBFB/2020\|EPI_ISL_573487\|20 | ···················· | ···················· | ····················· | ···················· | ···················· | ···················· |
| L | 5147 | hCoV-19/England/QEUH-9AEC07/2020\|EPI_ISL_573488\|20 | ···················· | ···················· | ····················· | ···················· | ···················· | ···················· |
| L | 5148 | hCoV-19/England/QEUH-9AEC43/2020\|EPI_ISL_573489\|20 | ···················· | ···················· | ····················· | ···················· | ···················· | ···················· |
| L | 5149 | hCoV-19/England/QEUH-9AEC70/2020\|EPI_ISL_573490\|20 | ···················· | ···················· | ····················· | ···················· | ···················· | ···················· |
| L | 5150 | hCoV-19/England/QEUH-9AEC8F/2020\|EPI_ISL_573491\|20 | ···················· | ···················· | ····················· | ···················· | ···················· | ···················· |
| L | 5151 | hCoV-19/England/QEUH-9AEC9E/2020\|EPI_ISL_573492\|20 | ···················· | ···················· | ····················· | ···················· | ···················· | ···················· |
| L | 5152 | hCoV-19/England/QEUH-9AECAD/2020\|EPI_ISL_573493\|20 | ···················· | ···················· | ····················· | ···················· | ···················· | ···················· |
| L | 5153 | hCoV-19/England/QEUH-9AECCB/2020\|EPI_ISL_573494\|20 | ···················· | ···················· | ····················· | ···················· | ···················· | ···················· |
| L | 5154 | hCoV-19/England/QEUH-9AECE9/2020\|EPI_ISL_573495\|20 | ···················· | ···················· | ····················· | ···················· | ···················· | ···················· |
| L | 5155 | hCoV-19/England/QEUH-9AECF8/2020\|EPI_ISL_573496\|20 | ···················· | ···················· | ····················· | ···················· | ···················· | ···················· |
| L | 5156 | hCoV-19/England/QEUH-9AED04/2020\|EPI_ISL_573497\|20 | ···················· | ···················· | ····················· | ···················· | ···················· | ···················· |
| L | 5157 | hCoV-19/England/204020547/2020\|EPI_ISL_574700\|2020 | ···················· | ·A·················· | ····················· | ···················· | ···················· | ··A················· |
| L | 5158 | hCoV-19/England/204020548/2020\|EPI_ISL_574701\|2020 | ···················· | ·A·················· | ····················· | ···················· | ···················· | ··A················· |
| L | 5159 | hCoV-19/England/204020549/2020\|EPI_ISL_574702\|2020 | ···················· | ·A·················· | ····················· | ···················· | ···················· | ··A················· |
| L | 5160 | hCoV-19/England/204020550/2020\|EPI_ISL_574703\|2020 | ···················· | ·A·················· | ····················· | ···················· | ···················· | ··A················· |
| L | 5161 | hCoV-19/England/204020551/2020\|EPI_ISL_574704\|2020 | ···················· | ·A·················· | ····················· | ···················· | ···················· | ··A················· |
| L | 5162 | hCoV-19/Spain/RI-ISCIII-201573/2020\|EPI_ISL_578200 | ···················· | ···················· | ····················· | ···················· | ···················· | ···················· |
| L | 5163 | hCoV-19/Spain/RI-ISCIII-201574/2020\|EPI_ISL_578201 | ···················· | ···················· | ····················· | ···················· | ···················· | ···················· |
| L | 5164 | hCoV-19/Ireland/WD-NVRL-73IRL32924/2020\|EPI_ISL_57 | ···················· | ···················· | ····················· | ···················· | ···················· | ···················· |
| L | 5165 | hCoV-19/Ireland/CE-NVRL-72IRL34583/2020\|EPI_ISL_57 | ···················· | ·A·················· | ····················· | ···················· | ···················· | ··A················· |
| L | 5166 | hCoV-19/Ireland/CE-NVRL-73IRL22902/2020\|EPI_ISL_57 | ···················· | ···················· | ····················· | ···················· | ···················· | ···················· |
| L | 5167 | hCoV-19/Ireland/CE-NVRL-73IRL26235/2020\|EPI_ISL_57 | ···················· | ···················· | ····················· | ···················· | ···················· | ···················· |
| L | 5168 | hCoV-19/Ireland/CO-NVRL-72IRL32659/2020\|EPI_ISL_57 | ···················· | ···················· | ····················· | ···················· | ···················· | ···················· |
| L | 5169 | hCoV-19/Ireland/CO-NVRL-72IRL34461/2020\|EPI_ISL_57 | ···················· | ·A·················· | ····················· | ···················· | ···················· | ··A················· |
| L | 5170 | hCoV-19/Ireland/CO-NVRL-73IRL22588/2020\|EPI_ISL_57 | ···················· | ···················· | ····················· | ···················· | ···················· | ···················· |
| L | 5171 | hCoV-19/Ireland/CO-NVRL-73IRL22642/2020\|EPI_ISL_57 | ···················· | ···················· | ····················· | ···················· | ···················· | ···················· |
| L | 5172 | hCoV-19/Ireland/D-NVRL-72IRL33419/2020\|EPI_ISL_578 | ···················· | ···················· | ····················· | ···················· | ···················· | ···················· |
| L | 5173 | hCoV-19/Ireland/D-NVRL-73IRL27757/2020\|EPI_ISL_578 | ···················· | ···················· | ····················· | ···················· | ···················· | ···················· |
| L | 5174 | hCoV-19/Ireland/D-NVRL-73IRL27788/2020\|EPI_ISL_578 | ···················· | ···················· | ····················· | ···················· | ···················· | ···················· |
| L | 5175 | hCoV-19/Ireland/D-NVRL-73IRL34820/2020\|EPI_ISL_578 | ···················· | ···················· | ····················· | ···················· | ···················· | ···················· |
| L | 5176 | hCoV-19/Ireland/D-NVRL-73IRL37632/2020\|EPI_ISL_578 | ···················· | ···················· | ····················· | ···················· | ···················· | ···················· |
| L | 5177 | hCoV-19/Ireland/D-NVRL-73IRL38296/2020\|EPI_ISL_578 | ···················· | ···················· | ····················· | ···················· | ···················· | ···················· |
| L | 5178 | hCoV-19/Ireland/D-NVRL-73IRL43808/2020\|EPI_ISL_578 | ···················· | ···················· | ····················· | ···················· | ···················· | ···················· |
| L | 5179 | hCoV-19/Ireland/G-NVRL-20IRL40646-2/2020\|EPI_ISL_5 | ···················· | ·A·················· | ····················· | ···················· | ···················· | ··A················· |
| L | 5181 | hCoV-19/Ireland/G-NVRL-73IRL20450/2020\|EPI_ISL_578 | ···················· | ···················· | ····················· | ···················· | ···················· | ···················· |
| L | 5182 | hCoV-19/Ireland/G-NVRL-73IRL24800/2020\|EPI_ISL_578 | ···················· | ···················· | ····················· | ···················· | ···················· | ···················· |
| L | 5183 | hCoV-19/Ireland/G-NVRL-73IRL28112/2020\|EPI_ISL_578 | ···················· | ···················· | ····················· | ···················· | ···················· | ···················· |
| L | 5184 | hCoV-19/Ireland/G-NVRL-73IRL28287/2020\|EPI_ISL_578 | ···················· | ···················· | ····················· | ···················· | ···················· | ···················· |
| L | 5185 | hCoV-19/Ireland/KE-NVRL-72IRL24485/2020\|EPI_ISL_57 | ···················· | ···················· | ····················· | ···················· | ···················· | ···················· |
| L | 5186 | hCoV-19/Ireland/KE-NVRL-72IRL37291/2020\|EPI_ISL_57 | ···················· | ···················· | ····················· | ···················· | ···················· | ···················· |
| L | 5187 | hCoV-19/Ireland/KE-NVRL-72IRL76432/2020\|EPI_ISL_57 | ···················· | ···················· | ····················· | ···················· | ···················· | ···················· |
| L | 5188 | hCoV-19/Ireland/KE-NVRL-73IRL13506/2020\|EPI_ISL_57 | ···················· | ···················· | ····················· | ···················· | ···················· | ···················· |
| L | 5189 | hCoV-19/Ireland/KE-NVRL-73IRL13510/2020\|EPI_ISL_57 | ···················· | ···················· | ····················· | ···················· | ···················· | ···················· |
| L | 5190 | hCoV-19/Ireland/KE-NVRL-73IRL13512/2020\|EPI_ISL_57 | ···················· | ···················· | ····················· | ···················· | ···················· | ···················· |
| L | 5191 | hCoV-19/Ireland/KE-NVRL-73IRL13514/2020\|EPI_ISL_57 | ···················· | ···················· | ····················· | ···················· | ···················· | ···················· |
| L | 5192 | hCoV-19/Ireland/KE-NVRL-73IRL13515/2020\|EPI_ISL_57 | ···················· | ···················· | ····················· | ···················· | ···················· | ···················· |
| L | 5193 | hCoV-19/Ireland/KE-NVRL-73IRL13837/2020\|EPI_ISL_57 | ···················· | ···················· | ····················· | ···················· | ···················· | ···················· |
| L | 5194 | hCoV-19/Ireland/KE-NVRL-73IRL23016/2020\|EPI_ISL_57 | ···················· | ···················· | ····················· | ···················· | ···················· | ···················· |
| L | 5195 | hCoV-19/Ireland/KE-NVRL-73IRL25898/2020\|EPI_ISL_57 | ···················· | ···················· | ····················· | ···················· | ···················· | ···················· |
| L | 5196 | hCoV-19/Ireland/KE-NVRL-73IRL25939/2020\|EPI_ISL_57 | ···················· | ···················· | ····················· | ···················· | ···················· | ···················· |
| L | 5197 | hCoV-19/Ireland/KE-NVRL-73IRL25948/2020\|EPI_ISL_57 | ···················· | ···················· | ····················· | ···················· | ···················· | ···················· |
| L | 5198 | hCoV-19/Ireland/KE-NVRL-73IRL25972/2020\|EPI_ISL_57 | ···················· | ···················· | ····················· | ···················· | ···················· | ···················· |
| L | 5199 | hCoV-19/Ireland/KE-NVRL-73IRL28838/2020\|EPI_ISL_57 | ···················· | ···················· | ····················· | ···················· | ···················· | ···················· |
| L | 5200 | hCoV-19/Ireland/KE-NVRL-73IRL29421/2020\|EPI_ISL_57 | ···················· | ···················· | ····················· | ···················· | ···················· | ···················· |
| L | 5201 | hCoV-19/Ireland/KE-NVRL-73IRL29560/2020\|EPI_ISL_57 | ···················· | ···················· | ····················· | ···················· | ···················· | ···················· |
| L | 5202 | hCoV-19/Ireland/KE-NVRL-73IRL29633/2020\|EPI_ISL_57 | ···················· | ···················· | ····················· | ···················· | ···················· | ···················· |
| L | 5203 | hCoV-19/New Zealand/20VR3482/2020\|EPI_ISL_579500\|2 | ···················· | ···················· | ····················· | ···················· | ···················· | ···················· |
| L | 5204 | hCoV-19/New Zealand/20VR3483/2020\|EPI_ISL_579501\|2 | ···················· | ···················· | ····················· | ···················· | ···················· | ···················· |
| L | 5205 | hCoV-19/New Zealand/20VR3489/2020\|EPI_ISL_579503\|2 | ···················· | ···················· | ····················· | ···················· | ···················· | ···················· |
| L | 5206 | hCoV-19/New Zealand/20VR3490/2020\|EPI_ISL_579504\|2 | ···················· | ···················· | ····················· | ···················· | ···················· | ···················· |
| L | 5207 | hCoV-19/New Zealand/20VR3491/2020\|EPI_ISL_579505\|2 | ···················· | ···················· | ····················· | ···················· | ···················· | ···················· |
| L | 5208 | hCoV-19/New Zealand/20VR3493/2020\|EPI_ISL_579506\|2 | ···················· | ···················· | ····················· | ···················· | ···················· | ···················· |
| L | 5213 | hCoV-19/New Zealand/20VR3766/2020\|EPI_ISL_579512\|2 | ···················· | ···················· | ····················· | ···················· | ···················· | ···················· |
| L | 5215 | hCoV-19/Canada/NS-NML-7/2020\|EPI_ISL_579515\|2020-0 | ···················· | ···················· | ····················· | ···················· | ···················· | ···················· |
| L | 5216 | hCoV-19/Canada/NS-NML-12/2020\|EPI_ISL_579516\|2020- | ···················· | ···················· | ····················· | ···················· | ···················· | ···················· |
| L | 5217 | hCoV-19/Canada/NS-NML-41/2020\|EPI_ISL_579517\|2020- | ···················· | ···················· | ····················· | ···················· | ···················· | ···················· |
| L | 5218 | hCoV-19/Canada/NS-NML-160/2020\|EPI_ISL_579530\|2020 | ···················· | ···················· | ····················· | ···················· | ···················· | ···················· |
| L | 5219 | hCoV-19/Canada/NS-NML-198/2020\|EPI_ISL_579534\|2020 | ···················· | ···················· | ····················· | ···················· | ···················· | ···················· |
| L | 5220 | hCoV-19/Canada/NS-NML-884/2020\|EPI_ISL_579537\|2020 | ···················· | ···················· | ····················· | ···················· | ···················· | ···················· |
| L | 5221 | hCoV-19/Canada/NS-NML-886/2020\|EPI_ISL_579538\|2020 | ···················· | ···················· | ····················· | ···················· | ···················· | ···················· |
| L | 5222 | hCoV-19/Canada/NS-NML-887/2020\|EPI_ISL_579539\|2020 | ···················· | ···················· | ····················· | ···················· | ···················· | ···················· |
| L | 5223 | hCoV-19/Canada/NS-NML-888/2020\|EPI_ISL_579540\|2020 | ···················· | ···················· | ····················· | ···················· | ···················· | ···················· |
| L | 5224 | hCoV-19/Canada/NS-NML-889/2020\|EPI_ISL_579541\|2020 | ···················· | ···················· | ····················· | ···················· | ···················· | ···················· |
| L | 5225 | hCoV-19/Canada/NS-NML-891/2020\|EPI_ISL_579542\|2020 | ···················· | ···················· | ····················· | ···················· | ···················· | ···················· |

| | | | | | | | | |
|---|---|---|---|---|---|---|---|---|
| L | 5226 | hCoV-19/Canada/NS-NML-893/2020\|EPI_ISL_579543\|2020 | ···················· | ···················· | ···················· | ···················· | ···················· | ···················· |
| L | 5227 | hCoV-19/Canada/NS-NML-895/2020\|EPI_ISL_579544\|2020 | ···················· | ···················· | ···················· | ···················· | ···················· | ···················· |
| L | 5228 | hCoV-19/Canada/NS-NML-896/2020\|EPI_ISL_579545\|2020 | ···················· | ···················· | ···················· | ···················· | ···················· | ···················· |
| L | 5229 | hCoV-19/Canada/NS-NML-897/2020\|EPI_ISL_579546\|2020 | ···T················ | ···················· | ···················· | ···················· | ···················· | ···················· |
| L | 5230 | hCoV-19/Canada/NS-NML-898/2020\|EPI_ISL_579547\|2020 | ···T················ | ···················· | ···················· | ···················· | ···················· | ···················· |
| L | 5231 | hCoV-19/Canada/NS-NML-899/2020\|EPI_ISL_579548\|2020 | ···················· | ···················· | ···················· | ···················· | ···················· | ···················· |
| L | 5232 | hCoV-19/Canada/NS-NML-900/2020\|EPI_ISL_579549\|2020 | ···················· | ···················· | ···················· | ···················· | ···················· | ···················· |
| L | 5233 | hCoV-19/Canada/NS-NML-901/2020\|EPI_ISL_579550\|2020 | ···················· | ···················· | ···················· | ···················· | ···················· | ···················· |
| L | 5234 | hCoV-19/Canada/NS-NML-902/2020\|EPI_ISL_579551\|2020 | ···················· | ···················· | ···················· | ···················· | ···················· | ···················· |
| L | 5235 | hCoV-19/Canada/NS-NML-903/2020\|EPI_ISL_579552\|2020 | ···················· | ···················· | ···················· | ···················· | ···················· | ···················· |
| L | 5236 | hCoV-19/Canada/NS-NML-904/2020\|EPI_ISL_579553\|2020 | ···················· | ···················· | ···················· | ···················· | ···················· | ···················· |
| L | 5237 | hCoV-19/Canada/NS-NML-905/2020\|EPI_ISL_579554\|2020 | ···················· | ···················· | ···················· | ···················· | ···················· | ···················· |
| L | 5238 | hCoV-19/Canada/NS-NML-906/2020\|EPI_ISL_579555\|2020 | ···················· | ···················· | ···················· | ···················· | ···················· | ···················· |
| L | 5239 | hCoV-19/Canada/NS-NML-907/2020\|EPI_ISL_579556\|2020 | ···················· | ···················· | ···················· | ···················· | ···················· | ···················· |
| L | 5240 | hCoV-19/Canada/NS-NML-908/2020\|EPI_ISL_579557\|2020 | ···················· | ···················· | ···················· | ···················· | ···················· | ···················· |
| L | 5241 | hCoV-19/Canada/NS-NML-909/2020\|EPI_ISL_579558\|2020 | ···················· | ···················· | ···················· | ···················· | ···················· | ···················· |
| L | 5242 | hCoV-19/Canada/NS-NML-910/2020\|EPI_ISL_579559\|2020 | ···················· | ···················· | ···················· | ···················· | ···················· | ···················· |
| L | 5243 | hCoV-19/Canada/NS-NML-917/2020\|EPI_ISL_579560\|2020 | ···················· | ···················· | ···················· | ···················· | ···················· | ···················· |
| L | 5244 | hCoV-19/Canada/NS-NML-918/2020\|EPI_ISL_579561\|2020 | ···················· | ···················· | ···················· | ···················· | ···················· | ···················· |
| L | 5245 | hCoV-19/Canada/NS-NML-919/2020\|EPI_ISL_579562\|2020 | ···················· | ···················· | ···················· | ···················· | ···················· | ···················· |
| L | 5246 | hCoV-19/Canada/NS-NML-920/2020\|EPI_ISL_579563\|2020 | ···················· | ···················· | ···················· | ···················· | ···················· | ···················· |
| L | 5247 | hCoV-19/Canada/NS-NML-922/2020\|EPI_ISL_579564\|2020 | ···················· | ···················· | ···················· | ···················· | ···················· | ···················· |
| L | 5248 | hCoV-19/Canada/NS-NML-924/2020\|EPI_ISL_579565\|2020 | ···················· | ···················· | ···················· | ···················· | ···················· | ···················· |
| L | 5249 | hCoV-19/Canada/NS-NML-926/2020\|EPI_ISL_579566\|2020 | ···················· | ·A·················· | ···················· | ···················· | ···················· | ···················· |
| L | 5250 | hCoV-19/Canada/NS-NML-927/2020\|EPI_ISL_579567\|2020 | ···················· | ···················· | ···················· | ···················· | ···················· | ···················· |
| L | 5251 | hCoV-19/Canada/NS-NML-928/2020\|EPI_ISL_579568\|2020 | ···················· | ···················· | ···················· | ···················· | ···················· | ···················· |
| L | 5252 | hCoV-19/Canada/NS-NML-929/2020\|EPI_ISL_579569\|2020 | ···················· | ···················· | ···················· | ···················· | ···················· | ···················· |
| L | 5253 | hCoV-19/Canada/NS-NML-930/2020\|EPI_ISL_579570\|2020 | ···················· | ···················· | ···················· | ···················· | ···················· | ···················· |
| L | 5254 | hCoV-19/Canada/NS-NML-939/2020\|EPI_ISL_579571\|2020 | ···················· | ···················· | ···················· | ···················· | ···················· | ···················· |
| L | 5255 | hCoV-19/Canada/NS-NML-940/2020\|EPI_ISL_579572\|2020 | ···················· | ···················· | ···················· | ···················· | ···················· | ···················· |
| L | 5256 | hCoV-19/Canada/NS-NML-941/2020\|EPI_ISL_579573\|2020 | ···················· | ···················· | ···················· | ···················· | ···················· | ···················· |
| L | 5257 | hCoV-19/Canada/NS-NML-943/2020\|EPI_ISL_579574\|2020 | ···················· | ···················· | ···················· | ···················· | ···················· | ···················· |
| L | 5258 | hCoV-19/Canada/NS-NML-944/2020\|EPI_ISL_579575\|2020 | ···················· | ···················· | ···················· | ···················· | ···················· | ···················· |
| L | 5259 | hCoV-19/Canada/NS-NML-946/2020\|EPI_ISL_579576\|2020 | ···················· | ···················· | ···················· | ···················· | ···················· | ···················· |
| L | 5260 | hCoV-19/Canada/NS-NML-947/2020\|EPI_ISL_579577\|2020 | ···················· | ···················· | ···················· | ···················· | ···················· | ···················· |
| L | 5261 | hCoV-19/Canada/NS-NML-949/2020\|EPI_ISL_579579\|2020 | ···················· | ···················· | ···················· | ···················· | ···················· | ···················· |
| L | 5262 | hCoV-19/Canada/NS-NML-950/2020\|EPI_ISL_579580\|2020 | ···················· | ···················· | ···················· | ···················· | ···················· | ···················· |
| L | 5263 | hCoV-19/Canada/NS-NML-951/2020\|EPI_ISL_579581\|2020 | ···················· | ···················· | ···················· | ···················· | ···················· | ···················· |
| L | 5264 | hCoV-19/Canada/NS-NML-952/2020\|EPI_ISL_579582\|2020 | ···················· | ···················· | ···················· | ···················· | ···················· | ···················· |
| L | 5265 | hCoV-19/Canada/NS-NML-953/2020\|EPI_ISL_579583\|2020 | ···················· | ···················· | ···················· | ···················· | ···················· | ···················· |
| L | 5266 | hCoV-19/Canada/NS-NML-957/2020\|EPI_ISL_579586\|2020 | ···················· | ···················· | ···················· | ···················· | ···················· | ···················· |
| L | 5267 | hCoV-19/England/ALDP-9E6F0A/2020\|EPI_ISL_580500\|20 | ···················· | ···················· | ···················· | ···················· | ···················· | ···················· |
| L | 5268 | hCoV-19/England/ALDP-9E6535/2020\|EPI_ISL_580501\|20 | ···················· | ···················· | ···················· | ···················· | ···················· | ···················· |
| L | 5269 | hCoV-19/England/CAMC-9EC3F9/2020\|EPI_ISL_580502\|20 | ···················· | ···················· | ···················· | ···················· | ······T············· | ···················· |
| L | 5270 | hCoV-19/England/ALDP-9E6A36/2020\|EPI_ISL_580503\|20 | ···················· | ···················· | ···················· | ···················· | ···················· | ···················· |
| L | 5271 | hCoV-19/England/ALDP-9E6FEC/2020\|EPI_ISL_580504\|20 | ···················· | ···················· | ···················· | ···················· | ···················· | ···················· |
| L | 5272 | hCoV-19/England/ALDP-9E6BF7/2020\|EPI_ISL_580505\|20 | ···················· | ···················· | ···················· | ···················· | ···················· | ···················· |
| L | 5273 | hCoV-19/England/ALDP-9E624D/2020\|EPI_ISL_580506\|20 | ···················· | ·A·················· | ···················· | ···················· | ···················· | ···················· |
| L | 5274 | hCoV-19/England/CAMC-9EC274/2020\|EPI_ISL_580508\|20 | ···················· | ···················· | ···················· | ···················· | ······T············· | ···················· |
| L | 5275 | hCoV-19/England/ALDP-9E6B8E/2020\|EPI_ISL_580510\|20 | ···················· | ·A·················· | ···················· | ···················· | ···················· | ···················· |
| L | 5276 | hCoV-19/England/CAMC-9EC669/2020\|EPI_ISL_580511\|20 | ···················· | ···················· | ···················· | ···················· | ···················· | ···················· |
| L | 5277 | hCoV-19/England/ALDP-9E678A/2020\|EPI_ISL_580512\|20 | ···················· | ···················· | ···················· | ···················· | ···················· | ···················· |
| L | 5278 | hCoV-19/England/ALDP-9E6C30/2020\|EPI_ISL_580513\|20 | ···················· | ···················· | ···················· | ···················· | ···················· | ···················· |
| L | 5279 | hCoV-19/England/ALDP-9E6966/2020\|EPI_ISL_580516\|20 | ···················· | ···················· | ···················· | ···················· | ···················· | ···················· |
| L | 5280 | hCoV-19/England/ALDP-9E668D/2020\|EPI_ISL_580517\|20 | ···················· | ···················· | ···················· | ···················· | ····T··············· | ···················· |
| L | 5281 | hCoV-19/England/ALDP-9E6957/2020\|EPI_ISL_580518\|20 | ···················· | ···················· | ···················· | ···················· | ······T············· | ···················· |
| L | 5282 | hCoV-19/England/ALDP-9E6F28/2020\|EPI_ISL_580519\|20 | ···················· | ···················· | ···················· | ···················· | ···················· | ···················· |
| L | 5283 | hCoV-19/England/ALDP-9E6CA9/2020\|EPI_ISL_580520\|20 | ···················· | ···················· | ···················· | ···················· | ···················· | ···················· |
| L | 5284 | hCoV-19/England/ALDP-9E73EF/2020\|EPI_ISL_580521\|20 | ···················· | ···················· | ···················· | ···················· | ···················· | ···················· |
| L | 5285 | hCoV-19/England/ALDP-9E6975/2020\|EPI_ISL_580522\|20 | ···················· | ···················· | ···················· | ···················· | ···················· | ···················· |
| L | 5286 | hCoV-19/England/ALDP-9E691B/2020\|EPI_ISL_580523\|20 | ···················· | ···················· | ···················· | ···················· | ···················· | ···················· |
| L | 5287 | hCoV-19/England/ALDP-9E67E4/2020\|EPI_ISL_580524\|20 | ···················· | ···················· | ···················· | ···················· | ···················· | ···················· |
| L | 5288 | hCoV-19/England/ALDP-9E62F2/2020\|EPI_ISL_580525\|20 | ···················· | ···················· | ···················· | ···················· | ···················· | ···················· |
| L | 5289 | hCoV-19/England/ALDP-9E6553/2020\|EPI_ISL_580526\|20 | ···················· | ···················· | ···················· | ···················· | ···················· | ···················· |
| L | 5290 | hCoV-19/England/ALDP-9E71E5/2020\|EPI_ISL_580527\|20 | ···················· | ···················· | ···················· | ···················· | ···················· | ···················· |
| L | 5291 | hCoV-19/England/ALDP-9E6FCE/2020\|EPI_ISL_580528\|20 | ···················· | ···················· | ···················· | ···················· | ···················· | ···················· |
| L | 5292 | hCoV-19/England/ALDP-9E733A/2020\|EPI_ISL_580529\|20 | ···················· | ···················· | ···················· | ···················· | ···················· | ···················· |
| L | 5293 | hCoV-19/England/ALDP-9E6AEB/2020\|EPI_ISL_580530\|20 | ···················· | ···················· | ···················· | ···················· | ···················· | ···················· |
| L | 5294 | hCoV-19/England/ALDP-9E6B9D/2020\|EPI_ISL_580531\|20 | ···················· | ···················· | ···················· | ···················· | ···················· | ···················· |
| L | 5295 | hCoV-19/England/CAMC-9EC3CC/2020\|EPI_ISL_580532\|20 | ···················· | ···················· | ···················· | ···················· | ···················· | ···················· |
| L | 5296 | hCoV-19/England/CAMC-9EC3BD/2020\|EPI_ISL_580533\|20 | ···················· | ···················· | ···················· | ···················· | ···················· | ···················· |
| L | 5297 | hCoV-19/England/ALDP-9E690C/2020\|EPI_ISL_580534\|20 | ···················· | ···················· | ···················· | ···················· | ···················· | ···················· |
| L | 5298 | hCoV-19/England/ALDP-9E7024/2020\|EPI_ISL_580535\|20 | ···················· | ···················· | ···················· | ···················· | ···················· | ···················· |
| L | 5299 | hCoV-19/England/CAMC-9EC21A/2020\|EPI_ISL_580536\|20 | ···················· | ···················· | ···················· | ···················· | ···················· | ···················· |
| L | 5300 | hCoV-19/England/ALDP-9E7455/2020\|EPI_ISL_580537\|20 | ···················· | ···················· | ···················· | ···················· | ···················· | ···················· |
| L | 5301 | hCoV-19/England/ALDP-9E714F/2020\|EPI_ISL_580538\|20 | ···················· | ···················· | ···················· | ···················· | ···················· | ···················· |
| L | 5302 | hCoV-19/England/ALDP-9E680F/2020\|EPI_ISL_580539\|20 | ···················· | ···················· | ···················· | ···················· | ···················· | ···················· |
| L | 5303 | hCoV-19/England/ALDP-9E6AFA/2020\|EPI_ISL_580540\|20 | ···················· | ···················· | ···················· | ···················· | ···················· | ···················· |
| L | 5304 | hCoV-19/England/ALDP-9E6C9A/2020\|EPI_ISL_580541\|20 | ···················· | ···················· | ···················· | ···················· | ···················· | ···················· |
| L | 5305 | hCoV-19/England/ALDP-9E69EE/2020\|EPI_ISL_580542\|20 | ···················· | ···················· | ···················· | ···················· | ···················· | ···················· |

| | | | | | | | | | |
|---|---|---|---|---|---|---|---|---|---|
| L | 5306 | hCoV-19/England/ALDP-9E6E49/2020\|EPI_ISL_580543\|20 | ···················· | ···················· | ···················· | ···················· | ···················· | ···················· | |
| L | 5307 | hCoV-19/England/ALDP-9E6BBB/2020\|EPI_ISL_580544\|20 | ···················· | ···················· | ···················· | ···················· | ···················· | ···················· | |
| L | 5308 | hCoV-19/England/ALDP-9E6632/2020\|EPI_ISL_580545\|20 | ···················· | ···················· | ···················· | ···················· | ···················· | ···················· | |
| L | 5309 | hCoV-19/England/ALDP-9E6A72/2020\|EPI_ISL_580546\|20 | ···················· | ···················· | ···················· | ···················· | ···················· | ···················· | |
| L | 5310 | hCoV-19/England/ALDP-9E67F3/2020\|EPI_ISL_580547\|20 | ···················· | ···················· | ···················· | ···················· | ···················· | ···················· | |
| L | 5311 | hCoV-19/England/ALDP-9E6ACD/2020\|EPI_ISL_580548\|20 | ···················· | ···················· | ···················· | ···················· | ···················· | ···················· | |
| L | 5312 | hCoV-19/England/ALDP-9E6061/2020\|EPI_ISL_580551\|20 | ···················· | ···················· | ···················· | ···················· | ···················· | ···················· | |
| L | 5313 | hCoV-19/England/ALDP-9E6FA0/2020\|EPI_ISL_580552\|20 | ···················· | ·A·················· | ···················· | ···················· | ···················· | ···················· | |
| L | 5314 | hCoV-19/England/ALDP-9E6993/2020\|EPI_ISL_580553\|20 | ···················· | ···················· | ···················· | ···················· | ···················· | ···················· | |
| L | 5315 | hCoV-19/England/ALDP-9E6F37/2020\|EPI_ISL_580555\|20 | ···················· | ···················· | ···················· | ···················· | ···················· | ···················· | |
| L | 5316 | hCoV-19/England/CAMC-9EC57B/2020\|EPI_ISL_580557\|20 | ···················· | ···················· | ···················· | ···················· | ···················· | ···················· | |
| L | 5317 | hCoV-19/England/ALDP-9E69DF/2020\|EPI_ISL_580558\|20 | ···················· | ···················· | ···················· | ···················· | ···················· | ···················· | |
| L | 5318 | hCoV-19/England/CAMC-9EC5D5/2020\|EPI_ISL_580559\|20 | ···················· | ···················· | ···················· | ···················· | ···················· | ···················· | |
| L | 5319 | hCoV-19/England/ALDP-9E6B42/2020\|EPI_ISL_580561\|20 | ···················· | ···················· | ···················· | ···················· | ···················· | ···················· | |
| L | 5320 | hCoV-19/England/CAMC-9EC687/2020\|EPI_ISL_580564\|20 | ···················· | ···················· | ···················· | ···················· | ···················· | ···················· | |
| L | 5321 | hCoV-19/England/CAMC-9EC49C/2020\|EPI_ISL_580565\|20 | ···················· | ···················· | ···················· | ···················· | ···················· | ···················· | |
| L | 5322 | hCoV-19/England/ALDP-9E6210/2020\|EPI_ISL_580566\|20 | ···················· | ···················· | ···················· | ···················· | ···················· | ···················· | |
| L | 5323 | hCoV-19/England/ALDP-9E66BA/2020\|EPI_ISL_580568\|20 | ···················· | ···················· | ···················· | ···················· | ···················· | ···················· | |
| L | 5324 | hCoV-19/England/CAMC-9EC55D/2020\|EPI_ISL_580569\|20 | ···················· | ···················· | ···················· | ···················· | ···················· | ···················· | |
| L | 5325 | hCoV-19/England/ALDP-9E7297/2020\|EPI_ISL_580570\|20 | ···················· | ·A·················· | ···················· | ···················· | ···················· | ··A················· | |
| L | 5326 | hCoV-19/England/ALDP-9E7060/2020\|EPI_ISL_580571\|20 | ···················· | ···················· | ···················· | ···················· | ···················· | ···················· | |
| L | 5327 | hCoV-19/England/ALDP-9E6C6D/2020\|EPI_ISL_580572\|20 | ···················· | ···················· | ···················· | ···················· | ···················· | ···················· | |
| L | 5328 | hCoV-19/England/CAMC-9EC775/2020\|EPI_ISL_580573\|20 | ···················· | ·A·················· | ···················· | ···················· | ···················· | ··A················· | |
| L | 5329 | hCoV-19/England/ALDP-9E676C/2020\|EPI_ISL_580574\|20 | ···················· | ·A·················· | ···················· | ···················· | ···················· | ··A················· | |
| L | 5330 | hCoV-19/England/ALDP-9E630E/2020\|EPI_ISL_580575\|20 | ···················· | ···················· | ···················· | ···················· | ···················· | ···················· | |
| L | 5331 | hCoV-19/England/CAMC-9EC48D/2020\|EPI_ISL_580576\|20 | ···················· | ···················· | ···················· | ···················· | ···················· | ···················· | |
| L | 5332 | hCoV-19/England/CAMC-9EC2DE/2020\|EPI_ISL_580577\|20 | ···················· | ···················· | ···················· | ···················· | ···················· | ···················· | |
| L | 5333 | hCoV-19/England/ALDP-9E6C8B/2020\|EPI_ISL_580578\|20 | ···················· | ···················· | ···················· | ···················· | ···················· | ···················· | |
| L | 5334 | hCoV-19/England/CAMC-9EC317/2020\|EPI_ISL_580579\|20 | ···················· | ···················· | ···················· | ···················· | ···················· | ···················· | |
| L | 5335 | hCoV-19/England/ALDP-9E65F9/2020\|EPI_ISL_580580\|20 | ···················· | ···················· | ···················· | ···················· | ···················· | ···················· | |
| L | 5336 | hCoV-19/England/ALDP-9E7385/2020\|EPI_ISL_580581\|20 | ···················· | ···················· | ···················· | ···················· | ···················· | ···················· | |
| L | 5337 | hCoV-19/Switzerland/BS-42210994/2020\|EPI_ISL_58181 | ···················· | ···················· | ···················· | ···················· | ···················· | ···················· | |
| L | 5338 | hCoV-19/Switzerland/AG-42216512/2020\|EPI_ISL_58181 | ···················· | ···················· | ···················· | ···················· | ·NN················· | ···················· | |
| L | 5339 | hCoV-19/Switzerland/BS-42217210/2020\|EPI_ISL_58182 | ···················· | ···················· | ···················· | ···················· | ···················· | ···················· | |
| L | 5341 | hCoV-19/Switzerland/BS-42217784/2020\|EPI_ISL_58182 | ···················· | ···················· | ···················· | ···················· | ···················· | ···················· | |
| L | 5342 | hCoV-19/Switzerland/BS-42217811/2020\|EPI_ISL_58182 | ···················· | ···················· | ···················· | ···················· | ···················· | ···················· | |
| L | 5343 | hCoV-19/Switzerland/BS-42218290/2020\|EPI_ISL_58183 | ···················· | ···················· | ···················· | ···················· | ···················· | ···················· | |
| L | 5344 | hCoV-19/Switzerland/BL-42218562/2020\|EPI_ISL_58183 | ···················· | ···················· | ···················· | ···················· | ···················· | ···················· | |
| L | 5345 | hCoV-19/Switzerland/BS-42218669/2020\|EPI_ISL_58183 | ···················· | ···················· | ···················· | ···················· | ·NN················· | ···················· | |
| L | 5346 | hCoV-19/Switzerland/BS-42218694/2020\|EPI_ISL_58183 | ···················· | ···················· | ···················· | ···················· | ···················· | ···················· | |
| L | 5347 | hCoV-19/Switzerland/BS-42218804/2020\|EPI_ISL_58183 | ···················· | ···················· | ···················· | ···················· | ···················· | ···················· | |
| L | 5348 | hCoV-19/Switzerland/BS-42218824/2020\|EPI_ISL_58183 | ···················· | ···················· | ···················· | ···················· | ···················· | ···················· | |
| L | 5349 | hCoV-19/Switzerland/BS-42218848/2020\|EPI_ISL_58184 | ···················· | ···················· | ···················· | ···················· | ···················· | ···················· | |
| L | 5350 | hCoV-19/Switzerland/BS-42219008/2020\|EPI_ISL_58184 | ···················· | ···················· | ···················· | ···················· | ···················· | ···················· | |
| L | 5351 | hCoV-19/Switzerland/NW-42219445/2020\|EPI_ISL_58184 | ···················· | ···················· | ···················· | ···················· | ···················· | ···················· | |
| L | 5352 | hCoV-19/Switzerland/BL-42219951/2020\|EPI_ISL_58184 | ···················· | ···················· | ···················· | ···················· | ···················· | ···················· | |
| L | 5353 | hCoV-19/Switzerland/BL-42219953/2020\|EPI_ISL_58184 | ···················· | ···················· | ···················· | ···················· | ···················· | ···················· | |
| L | 5354 | hCoV-19/Switzerland/BS-42219995/2020\|EPI_ISL_58184 | ···················· | ···················· | ···················· | ···················· | ···················· | ···················· | |
| L | 5355 | hCoV-19/Switzerland/BS-42220148/2020\|EPI_ISL_58185 | ···················· | ···················· | ···················· | ···················· | ···················· | ···················· | |
| L | 5356 | hCoV-19/Switzerland/BS-42220222/2020\|EPI_ISL_58185 | ···················· | ···················· | ···················· | ···················· | ···················· | ···················· | |
| L | 5357 | hCoV-19/Switzerland/BS-42220223/2020\|EPI_ISL_58185 | ···················· | ···················· | ···················· | ···················· | ···················· | ···················· | |
| L | 5359 | hCoV-19/Switzerland/BS-42220855/2020\|EPI_ISL_58185 | ···················· | ···················· | ···················· | ···················· | ···················· | ···················· | |
| L | 5360 | hCoV-19/Switzerland/BS-42220889/2020\|EPI_ISL_58185 | ···················· | ···················· | ···················· | ···················· | ·NNN················ | ···················· | 1 |
| L | 5361 | hCoV-19/Switzerland/BL-42221712/2020\|EPI_ISL_58186 | ···················· | ···················· | ···················· | ···················· | ···················· | ···················· | |
| L | 5362 | hCoV-19/Switzerland/BS-42221749/2020\|EPI_ISL_58186 | ···················· | ···················· | ···················· | ···················· | ···················· | ···················· | |
| L | 5363 | hCoV-19/Switzerland/BL-65239904/2020\|EPI_ISL_58186 | ···················· | ···················· | ···················· | ···················· | ···················· | ···················· | |
| L | 5364 | hCoV-19/Switzerland/BL-65240056/2020\|EPI_ISL_58187 | ···················· | ···················· | ···················· | ···················· | ···················· | ···················· | |
| L | 5370 | hCoV-19/Switzerland/BS-42335263/2020\|EPI_ISL_58189 | ···················· | ···················· | ···················· | ···················· | ·NN················· | ···················· | |
| L | 5371 | hCoV-19/Switzerland/BS-42338923/2020\|EPI_ISL_58189 | ···················· | ···················· | ···················· | ···················· | ·NN················· | ···················· | |
| L | 5374 | hCoV-19/Switzerland/BS-42341001/2020\|EPI_ISL_58189 | ···················· | ···················· | ···················· | ···················· | ·NN················· | ···················· | |
| L | 5376 | hCoV-19/Switzerland/BS-42346079/2020\|EPI_ISL_58189 | ···················· | ···················· | ···················· | ···················· | ···················· | ···················· | |
| L | 5377 | hCoV-19/USA/CT-QDX-1404/2020\|EPI_ISL_572100\|2020-0 | ···················· | ···················· | ···················· | ···················· | ···················· | ···················· | |
| L | 5378 | hCoV-19/USA/CT-QDX-1405/2020\|EPI_ISL_572101\|2020-0 | ···················· | ···················· | ···················· | ···················· | ···················· | ···················· | |
| L | 5379 | hCoV-19/USA/CT-QDX-1406/2020\|EPI_ISL_572102\|2020-0 | ···················· | ···················· | ···················· | ···················· | ···················· | ···················· | |
| L | 5380 | hCoV-19/USA/CT-QDX-1407/2020\|EPI_ISL_572103\|2020-0 | ···················· | ···················· | ···················· | ···················· | ···················· | ···················· | |
| L | 5381 | hCoV-19/USA/MI-QDX-1439/2020\|EPI_ISL_572104\|2020-0 | ···················· | ···················· | ···················· | ···················· | ···················· | ···················· | |
| L | 5382 | hCoV-19/USA/MN-QDX-1316/2020\|EPI_ISL_572105\|2020-0 | ···················· | ···················· | ···················· | ···················· | ···················· | ···················· | |
| L | 5383 | hCoV-19/USA/MN-QDX-1317/2020\|EPI_ISL_572106\|2020-0 | ···················· | ···················· | ···················· | ···················· | ···················· | ···················· | |
| L | 5384 | hCoV-19/USA/MN-QDX-1318/2020\|EPI_ISL_572107\|2020-0 | ···················· | ···················· | ···················· | ···················· | ···················· | ···················· | |
| L | 5385 | hCoV-19/USA/MN-QDX-1319/2020\|EPI_ISL_572108\|2020-0 | ···················· | ···················· | ···················· | ···················· | ···················· | ···················· | |
| L | 5386 | hCoV-19/USA/MN-QDX-1320/2020\|EPI_ISL_572109\|2020-0 | ···················· | ···················· | ···················· | ···················· | ···················· | ···················· | |
| L | 5387 | hCoV-19/USA/MN-QDX-1321/2020\|EPI_ISL_572110\|2020-0 | ···················· | ···················· | ···················· | ···················· | ···················· | ···················· | |
| L | 5388 | hCoV-19/USA/MN-QDX-1322/2020\|EPI_ISL_572111\|2020-0 | ···················· | ···················· | ···················· | ···················· | ···················· | ···················· | |
| L | 5389 | hCoV-19/USA/MN-QDX-1323/2020\|EPI_ISL_572112\|2020-0 | ···················· | ···················· | ···················· | ···················· | ···················· | ···················· | |
| L | 5390 | hCoV-19/USA/MN-QDX-1324/2020\|EPI_ISL_572113\|2020-0 | ···················· | ···················· | ···················· | ···················· | ···················· | ···················· | |
| L | 5391 | hCoV-19/USA/MN-QDX-1325/2020\|EPI_ISL_572114\|2020-0 | ···················· | ···················· | ···················· | ···················· | ···················· | ···················· | |
| L | 5392 | hCoV-19/USA/MN-QDX-1326/2020\|EPI_ISL_572115\|2020-0 | ···················· | ···················· | ···················· | ···················· | ···················· | ···················· | |
| L | 5393 | hCoV-19/USA/MN-QDX-1327/2020\|EPI_ISL_572116\|2020-0 | ···················· | ···················· | ···················· | ···················· | ···················· | ···················· | |
| L | 5394 | hCoV-19/USA/MN-QDX-1328/2020\|EPI_ISL_572117\|2020-0 | ···················· | ···················· | ···················· | ···················· | ···················· | ···················· | |
| L | 5395 | hCoV-19/USA/MN-QDX-1329/2020\|EPI_ISL_572118\|2020-0 | ···················· | ···················· | ···················· | ···················· | ···················· | ···················· | |

```
L    5396 hCoV-19/USA/MN-QDX-1330/2020|EPI_ISL_572119|2020-0    ····················  ····················  ····················  ····················  ····················  ····················
L    5397 hCoV-19/USA/MN-QDX-1331/2020|EPI_ISL_572120|2020-0    ····················  ····················  ····················  ····················  ····················  ····················
L    5398 hCoV-19/USA/MN-QDX-1332/2020|EPI_ISL_572121|2020-0    ····················  ····················  ····················  ····················  ····················  ····················
L    5399 hCoV-19/USA/MN-QDX-1333/2020|EPI_ISL_572122|2020-0    ····················  ····················  ····················  ····················  ····················  ····················
L    5400 hCoV-19/USA/MN-QDX-1334/2020|EPI_ISL_572123|2020-0    ····················  ····················  ····················  ····················  ····················  ····················
L    5401 hCoV-19/USA/MN-QDX-1335/2020|EPI_ISL_572124|2020-0    ····················  ····················  ····················  ····················  ····················  ····················
L    5402 hCoV-19/USA/MN-QDX-1336/2020|EPI_ISL_572125|2020-0    ····················  ····················  ····················  ····················  ····················  ····················
L    5403 hCoV-19/USA/MN-QDX-1337/2020|EPI_ISL_572126|2020-0    ····················  ····················  ····················  ····················  ····················  ····················
L    5404 hCoV-19/USA/MN-QDX-1338/2020|EPI_ISL_572127|2020-0    ····················  ····················  ····················  ····················  ····················  ····················
L    5405 hCoV-19/USA/MN-QDX-1339/2020|EPI_ISL_572128|2020-0    ····················  ····················  ····················  ····················  ····················  ····················
L    5406 hCoV-19/USA/MN-QDX-1340/2020|EPI_ISL_572129|2020-0    ····················  ····················  ····················  ····················  ····················  ····················
L    5407 hCoV-19/USA/MN-QDX-1341/2020|EPI_ISL_572130|2020-0    ····················  ····················  ····················  ····················  ····················  ····················
L    5408 hCoV-19/USA/MN-QDX-1342/2020|EPI_ISL_572131|2020-0    ····················  ····················  ····················  ····················  ····················  ····················
L    5409 hCoV-19/USA/MN-QDX-1343/2020|EPI_ISL_572132|2020-0    ····················  ····················  ····················  ····················  ····················  ····················
L    5410 hCoV-19/USA/MN-QDX-1344/2020|EPI_ISL_572133|2020-0    ····················  ····················  ····················  ····················  ····················  ····················
L    5411 hCoV-19/USA/MN-QDX-1345/2020|EPI_ISL_572134|2020-0    ····················  ····················  ····················  ····················  ····················  ····················
L    5412 hCoV-19/USA/MN-QDX-1346/2020|EPI_ISL_572135|2020-0    ····················  ····················  ····················  ····················  ····················  ····················
L    5413 hCoV-19/USA/MN-QDX-1347/2020|EPI_ISL_572136|2020-0    ····················  ····················  ····················  ····················  ····················  ····················
L    5414 hCoV-19/USA/MN-QDX-1348/2020|EPI_ISL_572137|2020-0    ····················  ····················  ····················  ····················  ····················  ····················
L    5415 hCoV-19/USA/MN-QDX-1349/2020|EPI_ISL_572138|2020-0    ····················  ····················  ····················  ····················  ····················  ····················
L    5416 hCoV-19/USA/MN-QDX-1350/2020|EPI_ISL_572139|2020-0    ····················  ····················  ····················  ····················  ····················  ····················
L    5417 hCoV-19/USA/MN-QDX-1351/2020|EPI_ISL_572140|2020-0    ····················  ····················  ····················  ····················  ····················  ····················
L    5418 hCoV-19/USA/MN-QDX-1352/2020|EPI_ISL_572141|2020-0    ····················  ····················  ····················  ····················  ····················  ····················
L    5419 hCoV-19/USA/MN-QDX-1353/2020|EPI_ISL_572142|2020-0    ····················  ····················  ····················  ····················  ····················  ····················
L    5420 hCoV-19/USA/MN-QDX-1354/2020|EPI_ISL_572143|2020-0    ····················  ····················  ····················  ····················  ····················  ····················
L    5421 hCoV-19/USA/MN-QDX-1355/2020|EPI_ISL_572144|2020-0    ····················  ····················  ····················  ····················  ····················  ····················
L    5422 hCoV-19/USA/MN-QDX-1356/2020|EPI_ISL_572145|2020-0    ····················  ····················  ····················  ····················  ····················  ····················
L    5423 hCoV-19/USA/MN-QDX-1357/2020|EPI_ISL_572146|2020-0    ····················  ····················  ····················  ····················  ····················  ····················
L    5424 hCoV-19/USA/MN-QDX-1358/2020|EPI_ISL_572147|2020-0    ····················  ····················  ····················  ····················  ····················  ····················
L    5425 hCoV-19/USA/MN-QDX-1359/2020|EPI_ISL_572148|2020-0    ····················  ····················  ····················  ····················  ····················  ····················
L    5426 hCoV-19/USA/MN-QDX-1360/2020|EPI_ISL_572149|2020-0    ····················  ····················  ····················  ····················  ····················  ····················
L    5427 hCoV-19/USA/MN-QDX-1378/2020|EPI_ISL_572167|2020-0    ····················  ····················  ····················  ····················  ····················  ····················
L    5428 hCoV-19/USA/MN-QDX-1379/2020|EPI_ISL_572168|2020-0    ····················  ····················  ····················  ····················  ····················  ····················
L    5429 hCoV-19/USA/MN-QDX-1380/2020|EPI_ISL_572169|2020-0    ····················  ····················  ····················  ····················  ····················  ····················
L    5430 hCoV-19/USA/MN-QDX-1381/2020|EPI_ISL_572170|2020-0    ····················  ····················  ····················  ····················  ····················  ····················
L    5431 hCoV-19/USA/MN-QDX-1382/2020|EPI_ISL_572171|2020-0    ····················  ····················  ····················  ····················  ····················  ····················
L    5432 hCoV-19/USA/MN-QDX-1383/2020|EPI_ISL_572172|2020-0    ····················  ····················  ····················  ····················  ····················  ····················
L    5433 hCoV-19/USA/MN-QDX-1384/2020|EPI_ISL_572173|2020-0    ····················  ····················  ····················  ····················  ····················  ····················
L    5434 hCoV-19/USA/MN-QDX-1385/2020|EPI_ISL_572174|2020-0    ····················  ····················  ····················  ····················  ····················  ····················
L    5435 hCoV-19/USA/MN-QDX-1386/2020|EPI_ISL_572175|2020-0    ····················  ····················  ····················  ····················  ····················  ····················
L    5436 hCoV-19/USA/MN-QDX-1387/2020|EPI_ISL_572176|2020-0    ····················  ····················  ····················  ····················  ····················  ····················
L    5437 hCoV-19/USA/MN-QDX-1388/2020|EPI_ISL_572177|2020-0    ····················  ····················  ····················  ····················  ····················  ····················
L    5438 hCoV-19/USA/MN-QDX-1389/2020|EPI_ISL_572178|2020-0    ····················  ····················  ····················  ····················  ····················  ····················
L    5439 hCoV-19/USA/MN-QDX-1390/2020|EPI_ISL_572179|2020-0    ····················  ····················  ····················  ····················  ····················  ····················
L    5440 hCoV-19/USA/MO-QDX-1314/2020|EPI_ISL_572180|2020-0    ····················  ····················  ····················  ····················  ····················  ····················
L    5441 hCoV-19/USA/NV-QDX-1393/2020|EPI_ISL_572181|2020-0    ····················  ····················  ····················  ····················  ····················  ····················
L    5442 hCoV-19/USA/NV-QDX-1394/2020|EPI_ISL_572182|2020-0    ····················  ····················  ····················  ····················  ····················  ····················
L    5443 hCoV-19/USA/OH-QDX-1392/2020|EPI_ISL_572183|2020-0    ····················  ····················  ····················  ····················  ····················  ····················
L    5444 hCoV-19/USA/TX-QDX-1399/2020|EPI_ISL_572184|2020-0    ····················  ····················  ····················  ····················  ····················  ····················
L    5445 hCoV-19/USA/UT-QDX-1391/2020|EPI_ISL_572185|2020-0    ····················  ····················  ····················  ····················  ····················  ····················
L    5446 hCoV-19/USA/VA-DCLS-1452/2020|EPI_ISL_572195|2020-    ····················  ····················  ····················  ····················  ····················  ····················
L    5447 hCoV-19/USA/VA-DCLS-1453/2020|EPI_ISL_572196|2020-    ····················  ····················  ····················  ····················  ····················  ····················
L    5448 hCoV-19/USA/VA-DCLS-1454/2020|EPI_ISL_572197|2020-    ····················  ····················  ····················  ····················  ····················  ····················
L    5449 hCoV-19/USA/VA-DCLS-1455/2020|EPI_ISL_572198|2020-    ····················  ····················  ····················  ····················  ····················  ····················
L    5450 hCoV-19/USA/VA-DCLS-1456/2020|EPI_ISL_572199|2020-    ····················  ····················  ····················  ····················  ····················  ····················
L    5451 hCoV-19/England/NORT-2A4C9F/2020|EPI_ISL_573400|20    ····················  ····················  ····················  ····················  ····················  ····················
L    5452 hCoV-19/England/NORT-2A4CBD/2020|EPI_ISL_573401|20    ····················  ····················  ····················  ····················  ····················  ····················
L    5453 hCoV-19/England/NORT-2A4D05/2020|EPI_ISL_573402|20    ····················  ····················  ····················  ····················  ·····T··············  ····················
L    5454 hCoV-19/England/NORT-2A4D14/2020|EPI_ISL_573403|20    ····················  ····················  ····················  ····················  ····················  ····················
L    5455 hCoV-19/England/NORT-2A4D41/2020|EPI_ISL_573404|20    ····················  ····················  ····················  ····················  ····················  ····················
L    5456 hCoV-19/England/NORT-2A4D9C/2020|EPI_ISL_573405|20    ····················  ····················  ····················  ····················  ····················  ····················
L    5457 hCoV-19/England/NORT-2A4DAB/2020|EPI_ISL_573406|20    ····················  ····················  ····················  ····················  ····················  ····················
L    5458 hCoV-19/England/NORT-2A4DC9/2020|EPI_ISL_573407|20    ····················  ····················  ····················  ····················  ····················  ····················
L    5459 hCoV-19/England/NORT-2A4DF6/2020|EPI_ISL_573408|20    ····················  ····················  ····················  ····················  ····················  ····················
L    5460 hCoV-19/England/NORT-2A4E20/2020|EPI_ISL_573409|20    ····················  ····················  ····················  ····················  ····················  ····················
L    5461 hCoV-19/England/NORT-2A4E5D/2020|EPI_ISL_573410|20    ····················  ····················  ····················  ····················  ····················  ····················
L    5462 hCoV-19/England/NORT-2A4E7B/2020|EPI_ISL_573411|20    ····················  ····················  ····················  ····················  ····················  ····················
L    5463 hCoV-19/England/NORT-2A4E8A/2020|EPI_ISL_573412|20    ····················  ····················  ····················  ····················  ····················  ····················
L    5464 hCoV-19/England/NORT-2A4EA8/2020|EPI_ISL_573413|20    ····················  ····················  ····················  ····················  ····················  ····················
L    5469 hCoV-19/England/NOTT-1133D3/2020|EPI_ISL_573418|20    ····················  ·A··················  ····················  ····················  ····················  ···A················
L    5470 hCoV-19/England/NOTT-1133F1/2020|EPI_ISL_573419|20    ····················  ····················  ····················  ····················  ····················  ····················
L    5473 hCoV-19/England/NOTT-113713/2020|EPI_ISL_573422|20    ····················  ····················  ····················  ····················  ····················  ····················
L    5474 hCoV-19/England/NOTT-113722/2020|EPI_ISL_573423|20    ····················  ·A··················  ····················  ····················  ····················  ····················
L    5475 hCoV-19/England/NOTT-113731/2020|EPI_ISL_573424|20    ····················  ····················  ····················  ····················  ····················  ····················
L    5477 hCoV-19/England/NOTT-11377D/2020|EPI_ISL_573426|20    ····················  ····················  ····················  ····················  ····················  ····················
L    5478 hCoV-19/England/NOTT-11378C/2020|EPI_ISL_573427|20    ····················  ····················  ····················  ····················  ····················  ····················
L    5480 hCoV-19/England/NOTT-1137AA/2020|EPI_ISL_573429|20    ····················  ····················  ····················  ····················  ····················  ····················
L    5481 hCoV-19/England/NOTT-1137C8/2020|EPI_ISL_573430|20    ····················  ····················  ····················  ····················  ····················  ····················
L    5483 hCoV-19/England/NOTT-1137E6/2020|EPI_ISL_573432|20    ····················  ····················  ····················  ····················  ····················  ····················
L    5484 hCoV-19/England/NOTT-113801/2020|EPI_ISL_573433|20    ····················  ····················  ····················  ····················  ····················  ····················
```

| | | | | | | | |
|---|---|---|---|---|---|---|---|
| L | 5485 hCoV-19/England/NOTT-113810/2020\|EPI_ISL_573434\|20 | ···················· | ···················· | ····················· | ···················· | ···················· | ···················· |
| L | 5486 hCoV-19/England/NOTT-11382F/2020\|EPI_ISL_573435\|20 | ···················· | ···················· | ····················· | ···················· | ···················· | ···················· |
| L | 5487 hCoV-19/England/NOTT-11383E/2020\|EPI_ISL_573436\|20 | ···················· | ···················· | ····················· | ···················· | ···················· | ···················· |
| L | 5488 hCoV-19/England/NOTT-11385C/2020\|EPI_ISL_573437\|20 | ···················· | ···················· | ····················· | ···················· | ···················· | ···················· |
| L | 5489 hCoV-19/England/NOTT-11386B/2020\|EPI_ISL_573438\|20 | ···················· | ···················· | ····················· | ···················· | ···················· | ···················· |
| L | 5490 hCoV-19/England/NOTT-113898/2020\|EPI_ISL_573439\|20 | ···················· | ···················· | ····················· | ···················· | ···················· | ···················· |
| L | 5491 hCoV-19/England/NOTT-1138A7/2020\|EPI_ISL_573440\|20 | ···················· | ···················· | ····················· | ···················· | ···················· | ···················· |
| L | 5492 hCoV-19/England/NOTT-1138B6/2020\|EPI_ISL_573441\|20 | ···················· | ···················· | ····················· | ···················· | ···················· | ···················· |
| L | 5493 hCoV-19/England/NOTT-1138C5/2020\|EPI_ISL_573442\|20 | ···················· | ···················· | ····················· | ···················· | ···················· | ···················· |
| L | 5494 hCoV-19/England/NOTT-1138F2/2020\|EPI_ISL_573443\|20 | ···················· | ···················· | ····················· | ···················· | ···················· | ···················· |
| L | 5495 hCoV-19/England/NOTT-11390E/2020\|EPI_ISL_573444\|20 | ···················· | ···················· | ····················· | ···················· | ···················· | ···················· |
| L | 5496 hCoV-19/England/NOTT-11391D/2020\|EPI_ISL_573445\|20 | ···················· | ···················· | ····················· | ···················· | ···················· | ···················· |
| L | 5497 hCoV-19/England/NOTT-11392C/2020\|EPI_ISL_573446\|20 | ···················· | ···················· | ····················· | ···················· | ···················· | ···················· |
| L | 5499 hCoV-19/England/NOTT-113986/2020\|EPI_ISL_573448\|20 | ···················· | ···················· | ····················· | ···················· | ···················· | ···················· |
| L | 5500 hCoV-19/England/NOTT-1139FF/2020\|EPI_ISL_573449\|20 | ···················· | ···················· | ····················· | ···················· | ···················· | ···················· |
| L | 5501 hCoV-19/England/NOTT-113A47/2020\|EPI_ISL_573450\|20 | ···················· | ···················· | ····················· | ···················· | ···················· | ···················· |
| L | 5502 hCoV-19/England/NOTT-113A56/2020\|EPI_ISL_573451\|20 | ···················· | ···················· | ····················· | ···················· | ···················· | ···················· |
| L | 5503 hCoV-19/England/NOTT-113A74/2020\|EPI_ISL_573452\|20 | ···················· | ···················· | ····················· | ···················· | ···················· | ···················· |
| L | 5504 hCoV-19/England/NOTT-113A83/2020\|EPI_ISL_573453\|20 | ···················· | ···················· | ····················· | ···················· | ···················· | ···················· |
| L | 5505 hCoV-19/England/NOTT-113A92/2020\|EPI_ISL_573454\|20 | ···················· | ···················· | ····················· | ···················· | ···················· | ···················· |
| L | 5506 hCoV-19/England/NOTT-113AA1/2020\|EPI_ISL_573455\|20 | ···················· | ···················· | ····················· | ···················· | ···················· | ···················· |
| L | 5507 hCoV-19/England/NOTT-113ADE/2020\|EPI_ISL_573456\|20 | ···················· | ···················· | ····················· | ···················· | ···················· | ···················· |
| L | 5508 hCoV-19/England/NOTT-113AED/2020\|EPI_ISL_573457\|20 | ···················· | ···················· | ····················· | ···················· | ···················· | ···················· |
| L | 5509 hCoV-19/England/NOTT-113B08/2020\|EPI_ISL_573458\|20 | ···················· | ···················· | ····················· | ···················· | ···················· | ···················· |
| L | 5510 hCoV-19/England/NOTT-113B26/2020\|EPI_ISL_573459\|20 | ···················· | ···················· | ····················· | ···················· | ···················· | ···················· |
| L | 5511 hCoV-19/England/NOTT-113B35/2020\|EPI_ISL_573460\|20 | ···················· | ···················· | ····················· | ···················· | ···················· | ···················· |
| L | 5512 hCoV-19/England/NOTT-113B44/2020\|EPI_ISL_573461\|20 | ···················· | ···················· | ····················· | ···················· | ···················· | ···················· |
| L | 5513 hCoV-19/England/NOTT-113B71/2020\|EPI_ISL_573462\|20 | ···················· | ···················· | ····················· | ···················· | ···················· | ···················· |
| L | 5514 hCoV-19/England/NOTT-113B9F/2020\|EPI_ISL_573463\|20 | ···················· | ···················· | ····················· | ···················· | ···················· | ···················· |
| L | 5515 hCoV-19/England/NOTT-113BAE/2020\|EPI_ISL_573464\|20 | ···················· | ·A·················· | ····················· | ···················· | ···················· | ···A················ |
| L | 5516 hCoV-19/England/NOTT-113BBD/2020\|EPI_ISL_573465\|20 | ···················· | ···················· | ····················· | ···················· | ···················· | ···················· |
| L | 5517 hCoV-19/England/NOTT-113BCC/2020\|EPI_ISL_573466\|20 | ···················· | ·A·················· | ····················· | ···················· | ···················· | ···················· |
| L | 5518 hCoV-19/England/NOTT-113BF9/2020\|EPI_ISL_573467\|20 | ···················· | ···················· | ····················· | ···················· | ···················· | ···················· |
| L | 5519 hCoV-19/England/NOTT-113C14/2020\|EPI_ISL_573468\|20 | ···················· | ·A·················· | ····················· | ···················· | ···················· | ···A················ |
| L | 5520 hCoV-19/England/NOTT-113C32/2020\|EPI_ISL_573469\|20 | ···T················ | ···················· | ····················· | ···················· | ···················· | ···················· |
| L | 5521 hCoV-19/England/NOTT-113C7E/2020\|EPI_ISL_573470\|20 | ···················· | ···················· | ····················· | ···················· | ···················· | ···················· |
| L | 5522 hCoV-19/England/NOTT-113CAB/2020\|EPI_ISL_573471\|20 | ···················· | ···················· | ····················· | ···················· | ···················· | ···················· |
| L | 5523 hCoV-19/England/NOTT-113CBA/2020\|EPI_ISL_573472\|20 | ···················· | ···················· | ····················· | ···················· | ···················· | ···················· |
| L | 5524 hCoV-19/England/NOTT-113CC9/2020\|EPI_ISL_573473\|20 | ···················· | ···················· | ····················· | ···················· | ···················· | ···················· |
| L | 5525 hCoV-19/England/NOTT-113CE7/2020\|EPI_ISL_573474\|20 | ···················· | ···················· | ····················· | ···················· | ···················· | ···················· |
| L | 5526 hCoV-19/England/NOTT-113CF6/2020\|EPI_ISL_573475\|20 | ···················· | ···················· | ····················· | ···················· | ···················· | ···················· |
| L | 5527 hCoV-19/England/NOTT-113D11/2020\|EPI_ISL_573476\|20 | ···················· | ···················· | ····················· | ···················· | ···················· | ···················· |
| L | 5528 hCoV-19/England/NOTT-113D3F/2020\|EPI_ISL_573477\|20 | ···················· | ···················· | ····················· | ···················· | ···················· | ···················· |
| L | 5529 hCoV-19/England/QEUH-9AEB28/2020\|EPI_ISL_573479\|20 | ···················· | ···················· | ····················· | ···················· | ···················· | ···················· |
| L | 5530 hCoV-19/England/QEUH-9AED22/2020\|EPI_ISL_573498\|20 | ···················· | ···················· | ····················· | ···················· | ···················· | ···················· |
| L | 5531 hCoV-19/England/QEUH-9AED40/2020\|EPI_ISL_573499\|20 | ···················· | ···················· | ····················· | ···················· | ···················· | ···················· |
| L | 5532 hCoV-19/Netherlands/DR-EMC-11/2020\|EPI_ISL_574779\| | ···················· | ···················· | ····················· | ···················· | ···················· | ···················· |
| L | 5533 hCoV-19/Netherlands/DR-EMC-12/2020\|EPI_ISL_574780\| | ···················· | ···················· | ····················· | ···················· | ···················· | ···················· |
| L | 5534 hCoV-19/Netherlands/DR-EMC-13/2020\|EPI_ISL_574781\| | ···················· | ···················· | ····················· | ···················· | ···················· | ···················· |
| L | 5535 hCoV-19/Netherlands/NB-EMC-283/2020\|EPI_ISL_574782 | ···················· | ···················· | ····················· | ···················· | ···················· | ···················· |
| L | 5536 hCoV-19/Netherlands/NB-EMC-284/2020\|EPI_ISL_574783 | ···················· | ···················· | ····················· | ···················· | ···················· | ···················· |
| L | 5537 hCoV-19/Netherlands/NB-EMC-286/2020\|EPI_ISL_574784 | ···················· | ···················· | ····················· | ···················· | ···················· | ···················· |
| L | 5538 hCoV-19/Netherlands/NB-EMC-287/2020\|EPI_ISL_574785 | ···················· | ···················· | ····················· | ···················· | ···················· | ···················· |
| L | 5539 hCoV-19/Netherlands/ZH-EMC-474/2020\|EPI_ISL_574786 | ···················· | ···················· | ····················· | ···················· | ···················· | ···················· |
| L | 5540 hCoV-19/Netherlands/ZH-EMC-475/2020\|EPI_ISL_574787 | ···················· | ···················· | ····················· | ···················· | ······T············· | ···················· |
| L | 5541 hCoV-19/Netherlands/ZH-EMC-476/2020\|EPI_ISL_574788 | ···················· | ···················· | ····················· | ···················· | ···················· | ···················· |
| L | 5542 hCoV-19/Netherlands/ZH-EMC-477/2020\|EPI_ISL_574789 | ···················· | ···················· | ····················· | ···················· | ······T············· | ···················· |
| L | 5543 hCoV-19/Switzerland/BE-UNIBE-149-0641/2020\|EPI_ISL | ···················· | ···················· | ····················· | ···················· | ···················· | ···················· |
| L | 5544 hCoV-19/Switzerland/BE-UNIBE-150-5479/2020\|EPI_ISL | ···················· | ···················· | ····················· | ···················· | ···················· | ···················· |
| L | 5545 hCoV-19/Switzerland/BE-UNIBE-151-6539/2020\|EPI_ISL | ···················· | ···················· | ····················· | ···················· | ···················· | ···················· |
| L | 5546 hCoV-19/Switzerland/BE-UNIBE-152-7992/2020\|EPI_ISL | ···················· | ···················· | ····················· | ···················· | ···················· | ···················· |
| L | 5547 hCoV-19/Switzerland/BE-UNIBE-153-8040/2020\|EPI_ISL | ···················· | ···················· | ····················· | ···················· | ···················· | ···················· |
| L | 5548 hCoV-19/Switzerland/BE-UNIBE-154-8234/2020\|EPI_ISL | ············Y······· | ···················· | ····················· | ···················· | ···················· | ···················· |
| L | 5549 hCoV-19/Switzerland/BE-UNIBE-155-8269/2020\|EPI_ISL | ···················· | ···················· | ····················· | ···················· | ···················· | ···················· |
| L | 5550 hCoV-19/Switzerland/BE-UNIBE-156-8498/2020\|EPI_ISL | ···················· | ···················· | ····················· | ···················· | ···················· | ···················· |
| L | 5551 hCoV-19/Switzerland/BE-UNIBE-157-8499/2020\|EPI_ISL | ···················· | ···················· | ····················· | ···················· | ···················· | ···················· |
| L | 5552 hCoV-19/Switzerland/BE-UNIBE-159-9400/2020\|EPI_ISL | ···················· | ···················· | ····················· | ···················· | ···················· | ···················· |
| L | 5554 hCoV-19/USA/MI-UM-10033827292/2020\|EPI_ISL_578132\| | ···················· | ···················· | ····················· | ···················· | ···················· | ···················· |
| L | 5555 hCoV-19/USA/MI-UM-10033828725/2020\|EPI_ISL_578133\| | ···················· | ···················· | ····················· | ···················· | ···················· | ···················· |
| L | 5556 hCoV-19/USA/MI-UM-10033828729/2020\|EPI_ISL_578134\| | ···················· | ···················· | ····················· | ···················· | ···················· | ···················· |
| L | 5557 hCoV-19/USA/MI-UM-10033829306/2020\|EPI_ISL_578135\| | ···················· | ···················· | ····················· | ···················· | ···················· | ···················· |
| L | 5559 hCoV-19/USA/MI-UM-10033836752/2020\|EPI_ISL_578137\| | ···················· | ···················· | ····················· | ···················· | ···················· | ···················· |
| L | 5560 hCoV-19/USA/MI-UM-10033842393/2020\|EPI_ISL_578138\| | ···················· | ···················· | ····················· | ···················· | ···················· | ···················· |
| L | 5562 hCoV-19/USA/MI-UM-10033851497/2020\|EPI_ISL_578140\| | ···················· | ···················· | ····················· | ···················· | ···················· | ···················· |
| L | 5564 hCoV-19/USA/MI-UM-10033852483/2020\|EPI_ISL_578142\| | ···················· | ···················· | ····················· | ···················· | ···················· | ···················· |
| L | 5567 hCoV-19/USA/MI-UM-10033871586/2020\|EPI_ISL_578145\| | ···················· | ···················· | ····················· | ···················· | ···················· | ···················· |
| L | 5568 hCoV-19/USA/MI-UM-10033872085/2020\|EPI_ISL_578146\| | ···················· | ···················· | ····················· | ···················· | ···················· | ···················· |
| L | 5569 hCoV-19/USA/MI-UM-10033872219/2020\|EPI_ISL_578147\| | ···················· | ···················· | ····················· | ···················· | ···················· | ···················· |
| L | 5570 hCoV-19/USA/MI-UM-10033872707/2020\|EPI_ISL_578148\| | ···················· | ···················· | ····················· | ···················· | ···················· | ···················· |
| L | 5571 hCoV-19/USA/MI-UM-10033876035/2020\|EPI_ISL_578149\| | ···················· | ···················· | ····················· | ···················· | ···················· | ···················· |

| | | | | | | | | |
|---|---|---|---|---|---|---|---|---|
| L | 5573 hCoV-19/New Zealand/20VR3171/2020\|EPI_ISL_579461\|2 | ···················· | ···················· | ···················· | ···················· | ···················· | ···················· | |
| L | 5574 hCoV-19/New Zealand/20VR3172/2020\|EPI_ISL_579462\|2 | ···················· | ···················· | ···················· | ···················· | ···················· | ···················· | |
| L | 5575 hCoV-19/New Zealand/20VR3173/2020\|EPI_ISL_579463\|2 | ···················· | ···················· | ···················· | ···················· | ···················· | ···················· | |
| L | 5576 hCoV-19/New Zealand/20VR3175/2020\|EPI_ISL_579464\|2 | ···················· | ···················· | ···················· | ···················· | ···················· | ···················· | |
| L | 5577 hCoV-19/New Zealand/20VR3176/2020\|EPI_ISL_579465\|2 | ···················· | ···················· | ···················· | ···················· | ···················· | ···················· | |
| L | 5578 hCoV-19/New Zealand/20VR3177/2020\|EPI_ISL_579466\|2 | ···················· | ···················· | ···················· | ···················· | ···················· | ···················· | |
| L | 5580 hCoV-19/New Zealand/20VR3179/2020\|EPI_ISL_579468\|2 | ···················· | ···················· | ···················· | ···················· | ···················· | ···················· | |
| L | 5581 hCoV-19/New Zealand/20VR3181/2020\|EPI_ISL_579469\|2 | ···················· | ···················· | ···················· | ···················· | ···················· | ···················· | |
| L | 5582 hCoV-19/New Zealand/20VR3182/2020\|EPI_ISL_579470\|2 | ···················· | ···················· | ···················· | ···················· | ···················· | ···················· | |
| L | 5583 hCoV-19/New Zealand/20VR3183/2020\|EPI_ISL_579471\|2 | ···················· | ···················· | ···················· | ···················· | ···················· | ···················· | |
| L | 5584 hCoV-19/New Zealand/20VR3186/2020\|EPI_ISL_579474\|2 | ···················· | ···················· | ···················· | ···················· | ···················· | ···················· | |
| L | 5585 hCoV-19/New Zealand/20VR3187/2020\|EPI_ISL_579475\|2 | ···················· | ···················· | ···················· | ···················· | ···················· | ···················· | |
| L | 5586 hCoV-19/New Zealand/20VR3191/2020\|EPI_ISL_579478\|2 | ···················· | ···················· | ···················· | ···················· | ···················· | ···················· | |
| L | 5588 hCoV-19/Canada/NS-NML-1199/2020\|EPI_ISL_579600\|202 | ···················· | ···················· | ···················· | ···················· | ···················· | ···················· | |
| L | 5589 hCoV-19/Canada/NS-NML-1203/2020\|EPI_ISL_579604\|202 | ···················· | ···················· | ···················· | ···················· | ···················· | ···················· | |
| L | 5590 hCoV-19/Canada/NS-NML-1204/2020\|EPI_ISL_579605\|202 | ···················· | ···················· | ···················· | ···················· | ···················· | ···················· | |
| L | 5591 hCoV-19/Canada/NS-NML-1207/2020\|EPI_ISL_579607\|202 | ···················· | ···················· | ···················· | ···················· | ···················· | ···················· | |
| L | 5592 hCoV-19/Canada/NS-NML-1216/2020\|EPI_ISL_579613\|202 | ···················· | ···················A | ···················· | ···················· | ···················· | ···················· | 1 |
| L | 5593 hCoV-19/England/ALDP-9E6E94/2020\|EPI_ISL_580461\|20 | ···················· | ···················· | ···················· | ···················· | ···················· | ···················· | |
| L | 5594 hCoV-19/England/ALDP-9E708E/2020\|EPI_ISL_580462\|20 | ···················· | ···················· | ···················· | ···················· | ···················· | ···················· | |
| L | 5595 hCoV-19/England/ALDP-9E72C4/2020\|EPI_ISL_580463\|20 | ···················· | ···················· | ···················· | ···················· | ···················· | ···················· | |
| L | 5596 hCoV-19/England/ALDP-9E6DF1/2020\|EPI_ISL_580464\|20 | ···················· | ···················· | ···················· | ···················· | ···················· | ···················· | |
| L | 5597 hCoV-19/England/CAMC-9EC229/2020\|EPI_ISL_580465\|20 | ···················· | ···················· | ···················· | ···················· | ···················· | ···················· | |
| L | 5598 hCoV-19/England/ALDP-9E718B/2020\|EPI_ISL_580466\|20 | ···················· | ·A·················· | ···················· | ···················· | ···················· | ···A················ | |
| L | 5599 hCoV-19/England/CAMC-9EC47E/2020\|EPI_ISL_580467\|20 | ···················· | ···················· | ···················· | ···················· | ···················· | ···················· | |
| L | 5600 hCoV-19/England/ALDP-9E6E76/2020\|EPI_ISL_580468\|20 | ···················· | ···················· | ···················· | ···················· | ···················· | ···················· | |
| L | 5601 hCoV-19/England/ALDP-9E6623/2020\|EPI_ISL_580469\|20 | ···················· | ···················· | ···················· | ···················· | ···················· | ···················· | |
| L | 5602 hCoV-19/England/CAMC-9EC1D1/2020\|EPI_ISL_580471\|20 | ···················· | ···················· | ···················· | ···················· | ···················· | ···················· | |
| L | 5603 hCoV-19/England/ALDP-9E69C0/2020\|EPI_ISL_580472\|20 | ···················· | ···················· | ···················· | ···················· | ···················· | ···················· | |
| L | 5604 hCoV-19/England/ALDP-9E71F4/2020\|EPI_ISL_580473\|20 | ···················· | ···················· | ···················· | ···················· | ···················· | ···················· | |
| L | 5605 hCoV-19/England/ALDP-9E6A63/2020\|EPI_ISL_580475\|20 | ···················· | ···················· | ···················· | ···················· | ···················· | ···················· | |
| L | 5606 hCoV-19/England/ALDP-9E622F/2020\|EPI_ISL_580476\|20 | ···················· | ···················· | ···················· | ···················· | ···················· | ···················· | |
| L | 5607 hCoV-19/England/ALDP-9E67A8/2020\|EPI_ISL_580477\|20 | ···················· | ···················· | ···················· | ···················· | ···················· | ···················· | |
| L | 5608 hCoV-19/England/CAMC-9EC20B/2020\|EPI_ISL_580478\|20 | ···················· | ···················· | ···················· | ···················· | ···················· | ···················· | |
| L | 5609 hCoV-19/England/CAMC-9EC010/2020\|EPI_ISL_580479\|20 | ···················· | ···················· | ···················· | ···················· | ···················· | ···················· | |
| L | 5610 hCoV-19/Switzerland/BS-42207917/2020\|EPI_ISL_58179 | ···················· | ···················· | ···················· | ···················· | ···················· | ···················· | |
| L | 5611 hCoV-19/Switzerland/BS-42208062/2020\|EPI_ISL_58179 | ···················· | ···················· | ···················· | ···················· | ···················· | ···················· | |
| L | 5612 hCoV-19/Switzerland/BS-42208220/2020\|EPI_ISL_58179 | ···················· | ···················· | ···················· | ···················· | ···················· | ···················· | |
| L | 5613 hCoV-19/Switzerland/BS-42208621/2020\|EPI_ISL_58179 | ···················· | ···················· | ···················· | ···················· | ···················· | ···················· | |
| L | 5614 hCoV-19/Switzerland/BS-42350110/2020\|EPI_ISL_58190 | ···················· | ···················· | ···················· | ···················· | ···················· | ···················· | |
| L | 5620 hCoV-19/Switzerland/BL-42292327/2020\|EPI_ISL_58190 | ···················· | ···················· | ···················· | ···················· | ···················· | ···················· | |
| L | 5623 hCoV-19/Switzerland/BS-42241303/2020\|EPI_ISL_58191 | ···················· | ···················· | ···················· | ···················· | ···················· | ···················· | |
| L | 5624 hCoV-19/Switzerland/BS-42243953/2020\|EPI_ISL_58191 | ···················· | ···················· | ···················· | ···················· | ···················· | ···················· | |
| L | 5627 hCoV-19/Switzerland/BS-42252201/2020\|EPI_ISL_58192 | ···················· | ···················· | ···················· | ···················· | ···················· | ···················· | |
| L | 5628 hCoV-19/Switzerland/BL-42253363/2020\|EPI_ISL_58192 | ···················· | ···················· | ···················· | ···················· | ·NN················· | ···················· | |
| L | 5633 hCoV-19/England/MILK-973694/2020\|EPI_ISL_575900\|20 | ···················· | ···················· | ···················· | ···················· | ···················· | ···················· | |
| L | 5634 hCoV-19/Wales/MILK-9762D9/2020\|EPI_ISL_575901\|2020 | ···················· | ···················· | ···················· | ···················· | ···················· | ···················· | |
| L | 5635 hCoV-19/England/MILK-973324/2020\|EPI_ISL_575904\|20 | ···················· | ···················· | ···················· | ···················· | ···················· | ···················· | |
| L | 5636 hCoV-19/England/MILK-973306/2020\|EPI_ISL_575905\|20 | ···················· | ···················· | ···················· | ···················· | ···················· | ···················· | |
| L | 5637 hCoV-19/England/MILK-973421/2020\|EPI_ISL_575906\|20 | ···················· | ···················· | ···················· | ···················· | ···················· | ···················· | |
| L | 5638 hCoV-19/England/MILK-976628/2020\|EPI_ISL_575907\|20 | ···················· | ···················· | ···················· | ···················· | ···················· | ···················· | |
| L | 5639 hCoV-19/England/MILK-974648/2020\|EPI_ISL_575908\|20 | ···················· | ···················· | ···················· | ···················· | ···················· | ···················· | |
| L | 5640 hCoV-19/England/MILK-973667/2020\|EPI_ISL_575909\|20 | ···················· | ···················· | ···················· | ···················· | ···················· | ···················· | |
| L | 5641 hCoV-19/England/MILK-976682/2020\|EPI_ISL_575910\|20 | ···················· | ···················· | ···················· | ···················· | ···················· | ···················· | |
| L | 5642 hCoV-19/England/MILK-974675/2020\|EPI_ISL_575911\|20 | ···················· | ·A·················· | ···················· | ···················· | ···················· | ···A················ | |
| L | 5643 hCoV-19/England/MILK-974912/2020\|EPI_ISL_575913\|20 | ···················· | ···················· | ···················· | ···················· | ···················· | ···················· | |
| L | 5644 hCoV-19/England/MILK-97488E/2020\|EPI_ISL_575914\|20 | ···················· | ···················· | ···················· | ···················· | ···················· | ···················· | |
| L | 5645 hCoV-19/England/MILK-9766BF/2020\|EPI_ISL_575915\|20 | ···················· | ···················· | ···················· | ···················· | ······T············· | ···················· | |
| L | 5646 hCoV-19/England/MILK-97651C/2020\|EPI_ISL_575916\|20 | ···················· | ···················· | ···················· | ···················· | ···················· | ···················· | |
| L | 5647 hCoV-19/England/MILK-974A1F/2020\|EPI_ISL_575918\|20 | ···················· | ···················· | ···················· | ···················· | ···················· | ···················· | |
| L | 5648 hCoV-19/England/MILK-976901/2020\|EPI_ISL_575919\|20 | ···················· | ···················· | ···················· | ···················· | ···················· | ···················· | |
| L | 5649 hCoV-19/England/MILK-974806/2020\|EPI_ISL_575920\|20 | ···················· | ···················· | ···················· | ···················· | ···················· | ···················· | |
| L | 5650 hCoV-19/England/MILK-97453C/2020\|EPI_ISL_575921\|20 | ···················· | ···················· | ···················· | ···················· | ···················· | ···················· | |
| L | 5651 hCoV-19/England/MILK-97627F/2020\|EPI_ISL_575922\|20 | ···················· | ···················· | ···················· | ···················· | ···················· | ···················· | |
| L | 5652 hCoV-19/England/MILK-97348B/2020\|EPI_ISL_575923\|20 | ···················· | ···················· | ···················· | ···················· | ···················· | ···················· | |
| L | 5653 hCoV-19/England/MILK-97338E/2020\|EPI_ISL_575924\|20 | ···················· | ·A·················· | ···················· | ···················· | ···················· | ···················· | |
| L | 5654 hCoV-19/England/MILK-976567/2020\|EPI_ISL_575926\|20 | ···················· | ···················· | ···················· | ···················· | ···················· | ···················· | |
| L | 5655 hCoV-19/England/MILK-9733D9/2020\|EPI_ISL_575927\|20 | ···················· | ···················· | ···················· | ···················· | ···················· | ···················· | |
| L | 5656 hCoV-19/England/MILK-974A79/2020\|EPI_ISL_575929\|20 | ···················· | ···················· | ···················· | ···················· | ···················· | ···················· | |
| L | 5657 hCoV-19/USA/MI-UM-10033768108/2020\|EPI_ISL_578100\| | ···················· | ···················· | ···················· | ···················· | ···················· | ···················· | |
| L | 5658 hCoV-19/USA/MI-UM-10033768659/2020\|EPI_ISL_578101\| | ···················· | ···················· | ···················· | ···················· | ···················· | ···················· | |
| L | 5659 hCoV-19/USA/MI-UM-10033769080/2020\|EPI_ISL_578102\| | ···················· | ···················· | ···················· | ···················· | ···················· | ···················· | |
| L | 5660 hCoV-19/USA/MI-UM-10033774911/2020\|EPI_ISL_578103\| | ···················· | ···················· | ···················· | ···················· | ···················· | ···················· | |
| L | 5661 hCoV-19/USA/MI-UM-10033775516/2020\|EPI_ISL_578104\| | ···················· | ···················· | ···················· | ···················· | ···················· | ···················· | |
| L | 5662 hCoV-19/USA/MI-UM-10033776027/2020\|EPI_ISL_578105\| | ···················· | ···················· | ···················· | ···················· | ···················· | ···················· | |
| L | 5663 hCoV-19/USA/MI-UM-10033776047/2020\|EPI_ISL_578106\| | ···················· | ···················· | ···················· | ···················· | ···················· | ···················· | |
| L | 5665 hCoV-19/USA/MI-UM-10033798318/2020\|EPI_ISL_578122\| | ···················· | ···················· | ···················· | ···················· | ···················· | ···················· | |
| L | 5667 hCoV-19/USA/MI-UM-10033808660/2020\|EPI_ISL_578125\| | ···················· | ···················· | ···················· | ···················· | ···················· | ···················· | |
| L | 5668 hCoV-19/USA/MI-UM-10033809081/2020\|EPI_ISL_578126\| | ···················· | ···················· | ···················· | ···················· | ···················· | ···················· | |
| L | 5669 hCoV-19/USA/MI-UM-10033810376/2020\|EPI_ISL_578127\| | ···················· | ···················· | ···················· | ···················· | ···················· | ···················· | |

| | | | | | | | | |
|---|---|---|---|---|---|---|---|---|
| L | 5670 | hCoV-19/USA/MI-UM-10033811348/2020\|EPI_ISL_578128\| | .................... | .................... | .................... | .................... | .................... | .................... |
| L | 5671 | hCoV-19/USA/MI-UM-10033813745/2020\|EPI_ISL_578129\| | .................... | .................... | .................... | .................... | .................... | .................... |
| L | 5672 | hCoV-19/USA/MI-UM-10033818123/2020\|EPI_ISL_578131\| | .................... | .................... | .................... | .................... | .................... | .................... |
| L | 5673 | hCoV-19/USA/MI-UM-10033876044/2020\|EPI_ISL_578150\| | .................... | .................... | .................... | .................... | .................... | .................... |
| L | 5674 | hCoV-19/USA/MI-UM-10033882288/2020\|EPI_ISL_578151\| | .................... | .................... | .................... | .................... | .................... | .................... |
| L | 5678 | hCoV-19/USA/MI-UM-10033907638/2020\|EPI_ISL_578155\| | .................... | .................... | .................... | .................... | .................... | .................... |
| L | 5679 | hCoV-19/USA/MI-UM-10033916720/2020\|EPI_ISL_578156\| | .................... | .................... | .................... | .................... | .................... | .................... |
| L | 5683 | hCoV-19/USA/MI-UM-10033971920/2020\|EPI_ISL_578160\| | .................... | .................... | .................... | .................... | .................... | .................... |
| L | 5684 | hCoV-19/USA/MI-UM-10033973539/2020\|EPI_ISL_578161\| | .................... | .................... | .................... | .................... | .................... | .................... |
| L | 5685 | hCoV-19/USA/MI-UM-10033981968/2020\|EPI_ISL_578162\| | .................... | .................... | .................... | .................... | .................... | .................... |
| L | 5686 | hCoV-19/USA/MI-UM-10034004586/2020\|EPI_ISL_578163\| | .................... | .................... | .................... | .................... | .................... | .................... |
| L | 5687 | hCoV-19/USA/MI-UM-10034005736/2020\|EPI_ISL_578164\| | .................... | .................... | .................... | .................... | .................... | .................... |
| L | 5689 | hCoV-19/USA/MI-UM-10034010951/2020\|EPI_ISL_578166\| | .................... | .................... | .................... | .................... | .................... | .................... |
| L | 5691 | hCoV-19/India/WB-IICB-008/2020\|EPI_ISL_578168\|2020 | .................... | .................... | .................... | .................... | .................... | .................... |
| L | 5694 | hCoV-19/USA/MI-UM-10034135987/2020\|EPI_ISL_578171\| | .................... | .................... | .................... | .................... | .................... | .................... |
| L | 5697 | hCoV-19/USA/MI-UM-A00000000JU/2020\|EPI_ISL_578174\| | .................... | .................... | .................... | .................... | .................... | .................... |
| L | 5698 | hCoV-19/India/WB-IICB-009/2020\|EPI_ISL_578175\|2020 | .................... | .................... | .................... | .................... | .................... | .................... |
| L | 5699 | hCoV-19/France/ARA-08938-isl/2020\|EPI_ISL_578176\|2 | .................... | .................... | .................... | .................... | .................... | .................... |
| L | 5700 | hCoV-19/France/ARA-09112-isl/2020\|EPI_ISL_578177\|2 | .................... | .................... | .................... | .................... | .................... | .................... |
| L | 5701 | hCoV-19/India/WB-IICB-010/2020\|EPI_ISL_578178\|2020 | .................... | .................... | .................... | .................... | ...........T........ | .................... |
| L | 5702 | hCoV-19/India/WB-IICB-011/2020\|EPI_ISL_578179\|2020 | .................... | .................... | .................... | .................... | .................... | .................... |
| L | 5703 | hCoV-19/India/WB-IICB-012/2020\|EPI_ISL_578180\|2020 | .................... | .................... | .................... | .................... | .................... | .................... |
| L | 5704 | hCoV-19/India/WB-IICB-013/2020\|EPI_ISL_578181\|2020 | .................... | .................... | .................... | .................... | .................... | .................... |
| L | 5705 | hCoV-19/India/WB-IICB-014/2020\|EPI_ISL_578182\|2020 | .................... | .A.................. | .................... | .................... | .................... | .................... |
| L | 5706 | hCoV-19/India/WB-IICB-015/2020\|EPI_ISL_578183\|2020 | .................... | .................... | .................... | .................... | .................... | .................... |
| L | 5707 | hCoV-19/India/WB-IICB-016/2020\|EPI_ISL_578184\|2020 | .................... | .................... | .................... | .................... | .................... | .................... |
| L | 5709 | hCoV-19/Spain/AN-ISCIII-2013382/2020\|EPI_ISL_57818 | .................... | .................... | .................... | .................... | .................... | .................... |
| L | 5710 | hCoV-19/Spain/AN-ISCIII-2013676/2020\|EPI_ISL_57818 | .................... | .................... | .................... | .................... | .................... | .................... |
| L | 5711 | hCoV-19/Spain/AN-ISCIII-2013675/2020\|EPI_ISL_57818 | .................... | .................... | .................... | .................... | .................... | .................... |
| L | 5712 | hCoV-19/Spain/AR-ISCIII-201441/2020\|EPI_ISL_578189 | .................... | .................... | .................... | .................... | .................... | .................... |
| L | 5713 | hCoV-19/Spain/AR-ISCIII-201689/2020\|EPI_ISL_578190 | .................... | .................... | .................... | .................... | .................... | .................... |
| L | 5714 | hCoV-19/Spain/AR-ISCIII-201690/2020\|EPI_ISL_578191 | .................... | .................... | .................... | .................... | .................... | .................... |
| L | 5715 | hCoV-19/Spain/AR-ISCIII-201778/2020\|EPI_ISL_578192 | .................... | .................... | .................... | .................... | .................... | .................... |
| L | 5716 | hCoV-19/Spain/AR-ISCIII-201781/2020\|EPI_ISL_578193 | .................... | .................... | .................... | .................... | .................... | .................... |
| L | 5717 | hCoV-19/Spain/GA-ISCIII-201797/2020\|EPI_ISL_578194 | .................... | .................... | .................... | .................... | .................... | .................... |
| L | 5718 | hCoV-19/Spain/GA-ISCIII-201965/2020\|EPI_ISL_578195 | .................... | .................... | .................... | .................... | .................... | .................... |
| L | 5719 | hCoV-19/Spain/RI-ISCIII-201564/2020\|EPI_ISL_578196 | .................... | .................... | .................... | .................... | .................... | .................... |
| L | 5720 | hCoV-19/Spain/RI-ISCIII-201566/2020\|EPI_ISL_578197 | .................... | .................... | .................... | .................... | .................... | .................... |
| L | 5721 | hCoV-19/Spain/RI-ISCIII-201569/2020\|EPI_ISL_578198 | .................... | .................... | .................... | .................... | .................... | .................... |
| L | 5722 | hCoV-19/Spain/RI-ISCIII-201571/2020\|EPI_ISL_578199 | .................... | .................... | .................... | .................... | .................... | .................... |
| L | 5723 | hCoV-19/New Zealand/20VR3041/2020\|EPI_ISL_579400\|2 | .................... | .................... | .................... | .................... | .................... | .................... |
| L | 5727 | hCoV-19/New Zealand/20VR3065/2020\|EPI_ISL_579405\|2 | .................... | .................... | .................... | .................... | .................... | .................... |
| L | 5729 | hCoV-19/New Zealand/20VR3068/2020\|EPI_ISL_579407\|2 | .................... | .................... | .................... | .................... | .................... | .................... |
| L | 5734 | hCoV-19/New Zealand/20VR3075/2020\|EPI_ISL_579412\|2 | .................... | .................... | .................... | .................... | .................... | .................... |
| L | 5736 | hCoV-19/New Zealand/20VR3088/2020\|EPI_ISL_579415\|2 | .................... | .................... | .................... | .................... | .................... | .................... |
| L | 5738 | hCoV-19/New Zealand/20VR3093/2020\|EPI_ISL_579417\|2 | .................... | .................... | .................... | .................... | .................... | .................... |
| L | 5739 | hCoV-19/New Zealand/20VR3096/2020\|EPI_ISL_579418\|2 | .................... | .................... | .................... | .................... | .................... | .................... |
| L | 5740 | hCoV-19/New Zealand/20VR3098/2020\|EPI_ISL_579419\|2 | .................... | .................... | .................... | .................... | .................... | .................... |
| L | 5742 | hCoV-19/New Zealand/20VR3110/2020\|EPI_ISL_579421\|2 | .................... | .................... | .................... | .................... | .................... | .................... |
| L | 5744 | hCoV-19/New Zealand/20VR3116/2020\|EPI_ISL_579423\|2 | .................... | .................... | .................... | .................... | .................... | .................... |
| L | 5745 | hCoV-19/New Zealand/20VR3117/2020\|EPI_ISL_579424\|2 | .................... | .................... | .................... | .................... | .................... | .................... |
| L | 5746 | hCoV-19/New Zealand/20VR3119/2020\|EPI_ISL_579425\|2 | .................... | .................... | .................... | .................... | .................... | .................... |
| L | 5747 | hCoV-19/New Zealand/20VR3128/2020\|EPI_ISL_579426\|2 | .................... | .................... | .................... | .................... | .................... | .................... |
| L | 5750 | hCoV-19/New Zealand/20VR3138/2020\|EPI_ISL_579430\|2 | .................... | .................... | .................... | .................... | .................... | .................... |
| L | 5755 | hCoV-19/New Zealand/20VR3143/2020\|EPI_ISL_579435\|2 | .................... | .................... | .................... | .................... | .................... | .................... |
| L | 5759 | hCoV-19/New Zealand/20VR3163/2020\|EPI_ISL_579453\|2 | .................... | .................... | .................... | .................... | .................... | .................... |
| L | 5761 | hCoV-19/New Zealand/20VR3166/2020\|EPI_ISL_579456\|2 | .................... | .................... | .................... | .................... | .................... | .................... |
| L | 5763 | hCoV-19/New Zealand/20VR3168/2020\|EPI_ISL_579458\|2 | .................... | .................... | .................... | .................... | .................... | .................... |
| L | 5764 | hCoV-19/New Zealand/20VR3169/2020\|EPI_ISL_579459\|2 | .................... | .................... | .................... | .................... | .................... | .................... |
| L | 5765 | hCoV-19/New Zealand/20VR3194/2020\|EPI_ISL_579480\|2 | .................... | .................... | .................... | .................... | .................... | .................... |
| L | 5766 | hCoV-19/New Zealand/20VR3195/2020\|EPI_ISL_579481\|2 | .................... | .................... | .................... | .................... | .................... | .................... |
| L | 5767 | hCoV-19/New Zealand/20VR3196/2020\|EPI_ISL_579482\|2 | .................... | .................... | .................... | .................... | .................... | .................... |
| L | 5770 | hCoV-19/New Zealand/20VR3200/2020\|EPI_ISL_579485\|2 | .................... | .................... | .................... | .................... | .................... | .................... |
| L | 5772 | hCoV-19/New Zealand/20VR3202/2020\|EPI_ISL_579487\|2 | .................... | .................... | .................... | .................... | .................... | .................... |
| L | 5773 | hCoV-19/New Zealand/20VR3203/2020\|EPI_ISL_579488\|2 | .................... | .................... | .................... | .................... | .................... | .................... |
| L | 5777 | hCoV-19/New Zealand/20VR3209/2020\|EPI_ISL_579494\|2 | .................... | .................... | .................... | .................... | .................... | .................... |
| L | 5778 | hCoV-19/New Zealand/20VR3210/2020\|EPI_ISL_579495\|2 | .................... | .................... | .................... | .................... | .................... | .................... |
| L | 5779 | hCoV-19/New Zealand/20VR3476/2020\|EPI_ISL_579496\|2 | .................... | .................... | .................... | .................... | .................... | .................... |
| L | 5780 | hCoV-19/New Zealand/20VR3477/2020\|EPI_ISL_579497\|2 | .................... | .................... | .................... | .................... | .................... | .................... |
| L | 5781 | hCoV-19/New Zealand/20VR3480/2020\|EPI_ISL_579498\|2 | .................... | .................... | .................... | .................... | .................... | .................... |
| L | 5782 | hCoV-19/New Zealand/20VR3481/2020\|EPI_ISL_579499\|2 | .................... | .................... | .................... | .................... | .................... | .................... |
| L | 5783 | hCoV-19/England/CAMC-9EC70C/2020\|EPI_ISL_580400\|20 | .................... | .................... | .................... | .................... | .................... | .................... |
| L | 5784 | hCoV-19/England/ALDP-9E68A5/2020\|EPI_ISL_580402\|20 | .................... | .................... | .................... | .................... | .................... | .................... |
| L | 5785 | hCoV-19/England/ALDP-9E7367/2020\|EPI_ISL_580420\|20 | .................... | .................... | .................... | .................... | .................... | .................... |
| L | 5786 | hCoV-19/England/CAMC-9EC1FF/2020\|EPI_ISL_580421\|20 | .................... | .................... | .................... | .................... | .................... | .................... |
| L | 5787 | hCoV-19/England/ALDP-9E6D97/2020\|EPI_ISL_580422\|20 | .................... | .................... | .................... | .................... | .................... | .................... |
| L | 5788 | hCoV-19/England/ALDP-9E6948/2020\|EPI_ISL_580423\|20 | .................... | .................... | .................... | .................... | .................... | .................... |
| L | 5789 | hCoV-19/England/ALDP-9E6544/2020\|EPI_ISL_580424\|20 | .................... | .................... | .................... | .................... | .................... | .................... |
| L | 5790 | hCoV-19/England/ALDP-9E61F5/2020\|EPI_ISL_580425\|20 | .................... | .................... | .................... | .................... | .................... | .................... |
| L | 5791 | hCoV-19/England/ALDP-9E723D/2020\|EPI_ISL_580426\|20 | .................... | .................... | .................... | .................... | .................... | .................... |

| | | | | | | | | |
|---|---|---|---|---|---|---|---|---|
| L | 5792 hCoV-19/England/ALDP-9E6D5B/2020\|EPI_ISL_580427\|20 | ···················· | ···················· | ···················· | ···················· | ···················· | ···················· | |
| L | 5793 hCoV-19/England/ALDP-9E6DE2/2020\|EPI_ISL_580428\|20 | ···················· | ···················· | ···················· | ···················· | ···················· | ···················· | |
| L | 5794 hCoV-19/England/ALDP-9E71C7/2020\|EPI_ISL_580429\|20 | ···················· | ···················· | ···················· | ···················· | ···················· | ···················· | |
| L | 5795 hCoV-19/England/ALDP-9E68E1/2020\|EPI_ISL_580430\|20 | ···················· | ···················· | ···················· | ···················· | ···················· | ···················· | |
| L | 5796 hCoV-19/England/ALDP-9E631D/2020\|EPI_ISL_580431\|20 | ···················· | ···················· | ···················· | ···················· | ···················· | ···················· | |
| L | 5797 hCoV-19/England/ALDP-9E6878/2020\|EPI_ISL_580432\|20 | ···················· | ···················· | ···················· | ···················· | ···················· | ···················· | |
| L | 5798 hCoV-19/England/ALDP-9E7358/2020\|EPI_ISL_580433\|20 | ···················· | ···················· | ···················· | ···················· | ···················· | ···················· | |
| L | 5799 hCoV-19/England/ALDP-9E68D2/2020\|EPI_ISL_580434\|20 | ···················· | ···················· | ···················· | ···················· | ···················· | ···················· | |
| L | 5800 hCoV-19/England/CAMC-9EC4F6/2020\|EPI_ISL_580435\|20 | ···················· | ···················· | ···················· | ···················· | ···················· | ···················· | |
| L | 5801 hCoV-19/England/ALDP-9E62C5/2020\|EPI_ISL_580436\|20 | ···················· | ···················· | ···················· | ···················· | ···················· | ···················· | |
| L | 5802 hCoV-19/England/ALDP-9E683C/2020\|EPI_ISL_580437\|20 | ···················· | ·A·················· | ···················· | ···················· | ···················· | ··A················· | |
| L | 5803 hCoV-19/England/ALDP-9E6F55/2020\|EPI_ISL_580438\|20 | ···················· | ···················· | ···················· | ···················· | ···················· | ···················· | |
| L | 5804 hCoV-19/England/ALDP-9E7376/2020\|EPI_ISL_580439\|20 | ···················· | ···················· | ···················· | ···················· | ···················· | ···················· | |
| L | 5805 hCoV-19/England/ALDP-9E6E67/2020\|EPI_ISL_580440\|20 | ···················· | ···················· | ···················· | ···················· | ···················· | ···················· | |
| L | 5806 hCoV-19/England/ALDP-9E6298/2020\|EPI_ISL_580441\|20 | ···················· | ···················· | ···················· | ···················· | ···················· | ···················· | |
| L | 5807 hCoV-19/England/ALDP-9E71D6/2020\|EPI_ISL_580442\|20 | ···················· | ···················· | ···················· | ···················· | ···················· | ···················· | |
| L | 5808 hCoV-19/England/ALDP-9E66D8/2020\|EPI_ISL_580443\|20 | ···················· | ···················· | ···················· | ···················· | ···T················ | ···················· | |
| L | 5809 hCoV-19/England/CAMC-9EC2B0/2020\|EPI_ISL_580444\|20 | ···················· | ···················· | ···················· | ···················· | ···················· | ···················· | |
| L | 5810 hCoV-19/England/ALDP-9E707F/2020\|EPI_ISL_580445\|20 | ···················· | ···················· | ···················· | ···················· | ···················· | ···················· | |
| L | 5811 hCoV-19/England/ALDP-9E732B/2020\|EPI_ISL_580446\|20 | ···················· | ···················· | ···················· | ···················· | ···················· | ···················· | |
| L | 5812 hCoV-19/England/ALDP-9E685A/2020\|EPI_ISL_580447\|20 | ···················· | ·A·················· | ···················· | ···················· | ···················· | ···················· | |
| L | 5813 hCoV-19/England/ALDP-9E722E/2020\|EPI_ISL_580448\|20 | ···················· | ···················· | ···················· | ···················· | ···················· | ···················· | |
| L | 5814 hCoV-19/England/CAMC-9EC159/2020\|EPI_ISL_580449\|20 | ···················· | ···················· | ···········T········ | ···················· | ···················· | ···················· | |
| L | 5815 hCoV-19/England/ALDP-9E6B15/2020\|EPI_ISL_580451\|20 | ···················· | ···················· | ···················· | ···················· | ···················· | ···················· | |
| L | 5816 hCoV-19/England/CAMC-9EC362/2020\|EPI_ISL_580453\|20 | ···················· | ·A·················· | ···················· | ···················· | ···················· | ···················· | |
| L | 5817 hCoV-19/England/ALDP-9E6B24/2020\|EPI_ISL_580454\|20 | ···················· | ···················· | ···················· | ···················· | ···················· | ···················· | |
| L | 5818 hCoV-19/England/ALDP-9E6EFE/2020\|EPI_ISL_580455\|20 | ···················· | ···················· | ···················· | ···················· | ···················· | ···················· | |
| L | 5819 hCoV-19/England/ALDP-9E6D6A/2020\|EPI_ISL_580457\|20 | ···················· | ···················· | ···················· | ···················· | ···················· | ···················· | |
| L | 5820 hCoV-19/England/ALDP-9E6C5E/2020\|EPI_ISL_580458\|20 | ···················· | ···················· | ···················· | ···················· | ···················· | ···················· | |
| L | 5821 hCoV-19/England/CAMC-9EC61E/2020\|EPI_ISL_580459\|20 | ···················· | ···················· | ···················· | ···················· | ···················· | ···················· | |
| L | 5822 hCoV-19/England/ALDP-9E724C/2020\|EPI_ISL_580460\|20 | ···················· | ···················· | ···················· | ···················· | ···················· | ···················· | |
| L | 5823 hCoV-19/England/CAMC-9EC53F/2020\|EPI_ISL_580480\|20 | ···················· | ···················· | ···················· | ···················· | ···················· | ···················· | |
| L | 5824 hCoV-19/England/CAMC-9EC04D/2020\|EPI_ISL_580481\|20 | ···················· | ···················· | ···················· | ···················· | ···················· | ···················· | |
| L | 5825 hCoV-19/England/ALDP-9E6386/2020\|EPI_ISL_580482\|20 | ··················Y· | ···················· | ···················· | ···················· | ···················· | ···················· | 1 |
| L | 5826 hCoV-19/England/ALDP-9E666F/2020\|EPI_ISL_580483\|20 | ···················· | ···················· | ···················· | ···················· | ···················· | ···················· | |
| L | 5827 hCoV-19/Wales/ALDP-9E6B51/2020\|EPI_ISL_580484\|2020 | ···················· | ···················· | ···················· | ···················· | ···················· | ···················· | |
| L | 5828 hCoV-19/England/CAMC-9EC195/2020\|EPI_ISL_580485\|20 | ···················· | ···················· | ···················· | ···················· | ···················· | ···················· | |
| L | 5829 hCoV-19/England/CAMC-9EC238/2020\|EPI_ISL_580486\|20 | ···················· | ···················· | ···················· | ···················· | ···················· | ···················· | |
| L | 5830 hCoV-19/England/ALDP-9E62E3/2020\|EPI_ISL_580487\|20 | ···················· | ···················· | ···················· | ···················· | ···················· | ···················· | |
| L | 5831 hCoV-19/England/ALDP-9E73B2/2020\|EPI_ISL_580488\|20 | ···················· | ···················· | ···················· | ···················· | ···················· | ···················· | |
| L | 5832 hCoV-19/England/ALDP-9E6070/2020\|EPI_ISL_580489\|20 | ···················· | ···················· | ···················· | ···················· | ···················· | ···················· | |
| L | 5833 hCoV-19/England/ALDP-9E6BAC/2020\|EPI_ISL_580490\|20 | ···················· | ···················· | ···················· | ···················· | ···················· | ···················· | |
| L | 5834 hCoV-19/England/ALDP-9E6711/2020\|EPI_ISL_580491\|20 | ···················· | ···················· | ···················· | ···················· | ···················· | ···················· | |
| L | 5835 hCoV-19/England/ALDP-9E65CC/2020\|EPI_ISL_580492\|20 | ···················· | ···················· | ···················· | ···················· | ···················· | ···················· | |
| L | 5836 hCoV-19/England/ALDP-9E6ABE/2020\|EPI_ISL_580493\|20 | ···················· | ···················· | ···················· | ···················· | ···················· | ···················· | |
| L | 5837 hCoV-19/England/CAMC-9EC2A1/2020\|EPI_ISL_580495\|20 | ···················· | ·A·················· | ···················· | ···················· | ···················· | ··A················· | |
| L | 5838 hCoV-19/England/ALDP-9E6939/2020\|EPI_ISL_580496\|20 | ···················· | ···················· | ···················· | ···················· | ···················· | ···················· | |
| L | 5839 hCoV-19/England/ALDP-9E6E85/2020\|EPI_ISL_580497\|20 | ···················· | ···················· | ···················· | ···················· | ···················· | ···················· | |
| L | 5840 hCoV-19/England/CAMC-9EC58A/2020\|EPI_ISL_580499\|20 | ···················· | ···················· | ···················· | ···················· | ···················· | ···················· | |
| L | 5841 hCoV-19/Switzerland/BS-42231705/2020\|EPI_ISL_58170 | ···················· | ···················· | ···················· | ···················· | ···················· | ···················· | |
| L | 5842 hCoV-19/Switzerland/BL-42232196/2020\|EPI_ISL_58170 | ···················· | ···················· | ···················· | ···················· | ···················· | ···················· | |
| L | 5843 hCoV-19/Switzerland/BS-42232220/2020\|EPI_ISL_58170 | ···················· | ·A·················· | ···················· | ···················· | ···················· | ···················· | |
| L | 5844 hCoV-19/Switzerland/BS-42232763/2020\|EPI_ISL_58170 | ···················· | ···················· | ···················· | ···················· | ···················· | ···················· | |
| L | 5845 hCoV-19/Switzerland/BS-42232843/2020\|EPI_ISL_58170 | ···················· | ···················· | ···················· | ···················· | ···················· | ···················· | |
| L | 5846 hCoV-19/Switzerland/BS-42232888/2020\|EPI_ISL_58170 | ···················· | ···················· | ···················· | ···················· | ···················· | ···················· | |
| L | 5847 hCoV-19/Switzerland/BS-42233707/2020\|EPI_ISL_58170 | ···················· | ·A·················· | ···················· | ···················· | ···················· | ···················· | |
| L | 5848 hCoV-19/Switzerland/BS-42233903/2020\|EPI_ISL_58170 | ···················· | ···················· | ···················· | ···················· | ···················· | ···················· | |
| L | 5849 hCoV-19/Switzerland/BS-42234673/2020\|EPI_ISL_58171 | ···················· | ···················· | ···················· | ···················· | ···················· | ···················· | |
| L | 5850 hCoV-19/Switzerland/BS-42236042/2020\|EPI_ISL_58171 | ···················· | ···················· | ···················· | ···················· | ···················· | ···················· | |
| L | 5851 hCoV-19/Switzerland/BL-42236095/2020\|EPI_ISL_58171 | ···················· | ···················· | ···················· | ···················· | ···················· | ···················· | |
| L | 5852 hCoV-19/Switzerland/SH-42239215/2020\|EPI_ISL_58171 | ···················· | ···················· | ···················· | ···················· | ···················· | ···················· | |
| L | 5853 hCoV-19/Switzerland/BS-42199846/2020\|EPI_ISL_58171 | ···················· | ···················· | ···················· | ···················· | ···················· | ···················· | |
| L | 5854 hCoV-19/Switzerland/BS-42202177/2020\|EPI_ISL_58171 | ···················· | ···················· | ···················· | ···················· | ···················· | ···················· | |
| L | 5855 hCoV-19/Switzerland/BS-42188723/2020\|EPI_ISL_58171 | ···················· | ···················· | ···················· | ···················· | ···················· | ···················· | |
| L | 5856 hCoV-19/Switzerland/BS-42203342/2020\|EPI_ISL_58172 | ···················· | ···················· | ···················· | ···················· | ···················· | ···················· | |
| L | 5857 hCoV-19/Switzerland/BS-42204639/2020\|EPI_ISL_58172 | ···················· | ···················· | ···················· | ···················· | ···················· | ···················· | |
| L | 5858 hCoV-19/Switzerland/BS-42204863/2020\|EPI_ISL_58173 | ···················· | ···················· | ···················· | ···················· | ···················· | ···················· | |
| L | 5859 hCoV-19/Switzerland/BS-42204879/2020\|EPI_ISL_58173 | ···················· | ···················· | ···················· | ···················· | ···················· | ···················· | |
| L | 5860 hCoV-19/Switzerland/BS-42204952/2020\|EPI_ISL_58173 | ···················· | ···················· | ···················· | ···················· | ···················· | ···················· | |
| L | 5863 hCoV-19/Switzerland/BS-42207595/2020\|EPI_ISL_58175 | ···················· | ···················· | ···················· | ···················· | ···················· | ···················· | |
| L | 5865 hCoV-19/Switzerland/BS-42207953/2020\|EPI_ISL_58175 | ···················· | ···················· | ···················· | ···················· | ···················· | ···················· | |
| L | 5867 hCoV-19/Switzerland/BS-42208050/2020\|EPI_ISL_58176 | ···················· | ···················· | ···················· | ···················· | ···················· | ···················· | |
| L | 5868 hCoV-19/Switzerland/BS-42208204/2020\|EPI_ISL_58176 | ···················· | ···················· | ···················· | ···················· | ···················· | ···················· | |
| L | 5871 hCoV-19/Switzerland/BL-42208281/2020\|EPI_ISL_58176 | ···················· | ···················· | ···················· | ···················· | ···················· | ···················· | |
| L | 5872 hCoV-19/Switzerland/BS-42208299/2020\|EPI_ISL_58176 | ···················· | ···················· | ···················· | ···················· | ···················· | ···················· | |
| L | 5874 hCoV-19/Switzerland/BS-42208691/2020\|EPI_ISL_58177 | ···················· | ···················· | ···················· | ···················· | ···················· | ···················· | |
| L | 5877 hCoV-19/Switzerland/BS-42209502/2020\|EPI_ISL_58177 | ···················· | ···················· | ···················· | ···················· | ·NNN················ | ···················· | 1 |
| L | 5879 hCoV-19/Switzerland/BS-42209534/2020\|EPI_ISL_58178 | ···················· | ···················· | ···················· | ···················· | ·NN················· | ···················· | |
| L | 5880 hCoV-19/Switzerland/BL-42209747/2020\|EPI_ISL_58178 | ···················· | ···················· | ···················· | ···················· | ···················· | ···················· | |
| L | 5881 hCoV-19/Switzerland/BS-42204899/2020\|EPI_ISL_58178 | ···················· | ···················· | ···················· | ···················· | ···················· | ···················· | |

| | | | | | | | | |
|---|---|---|---|---|---|---|---|---|
| L | 5882 hCoV-19/Switzerland/VD-42205435/2020\|EPI_ISL_58178 | .................... | .................... | .................... | .................... | .................... | .................... | |
| L | 5883 hCoV-19/Switzerland/BL-42206147/2020\|EPI_ISL_58178 | .................... | .................... | .................... | .................... | .................... | .................... | |
| L | 5884 hCoV-19/Switzerland/BS-42206315/2020\|EPI_ISL_58178 | .................... | ...................A | .................... | .................... | .................... | .................... | 1 |
| L | 5885 hCoV-19/Switzerland/BS-42206628/2020\|EPI_ISL_58178 | .................... | .................... | .................... | .................... | .................... | .................... | |
| L | 5886 hCoV-19/Switzerland/BS-42206629/2020\|EPI_ISL_58178 | .................... | .................... | .................... | .................... | .NN................. | .................... | |
| L | 5887 hCoV-19/Switzerland/BS-42207729/2020\|EPI_ISL_58178 | .................... | .................... | .................... | .................... | .................... | .................... | |
| L | 5888 hCoV-19/Switzerland/BL-42207903/2020\|EPI_ISL_58179 | .................... | .................... | .................... | .................... | .................... | .................... | |
| L | 5889 hCoV-19/USA/NY-QDX-1260/2020\|EPI_ISL_572024\|2020-0 | .................... | .................... | .................... | .................... | .................... | .................... | |
| L | 5890 hCoV-19/USA/NY-QDX-1261/2020\|EPI_ISL_572025\|2020-0 | .................... | .................... | .................... | .................... | .................... | .................... | |
| L | 5891 hCoV-19/USA/NY-QDX-1262/2020\|EPI_ISL_572026\|2020-0 | .................... | .................... | .................... | .................... | .................... | .................... | |
| L | 5892 hCoV-19/USA/NY-QDX-1263/2020\|EPI_ISL_572027\|2020-0 | .................... | .................... | .................... | .................... | .................... | .................... | |
| L | 5893 hCoV-19/USA/NY-QDX-1264/2020\|EPI_ISL_572028\|2020-0 | .................... | .................... | .................... | .................... | .................... | .................... | |
| L | 5894 hCoV-19/USA/NY-QDX-1265/2020\|EPI_ISL_572029\|2020-0 | .................... | .................... | .................... | .................... | .................... | .................... | |
| L | 5895 hCoV-19/USA/NY-QDX-1268/2020\|EPI_ISL_572031\|2020-0 | .................... | .................... | .................... | .................... | .................... | .................... | |
| L | 5896 hCoV-19/USA/OH-QDX-1190/2020\|EPI_ISL_572032\|2020-0 | .................... | .................... | .................... | .................... | .................... | .................... | |
| L | 5897 hCoV-19/USA/OR-QDX-1274/2020\|EPI_ISL_572033\|2020-0 | .................... | .................... | .................... | .................... | .................... | .................... | |
| L | 5898 hCoV-19/USA/PA-QDX-1191/2020\|EPI_ISL_572034\|2020-0 | .................... | .................... | .................... | .................... | .................... | .................... | |
| L | 5899 hCoV-19/USA/TX-QDX-1197/2020\|EPI_ISL_572035\|2020-0 | .................... | .................... | .................... | .................... | .................... | .................... | |
| L | 5900 hCoV-19/USA/TX-QDX-1198/2020\|EPI_ISL_572036\|2020-0 | .................... | .................... | .................... | .................... | .................... | .................... | |
| L | 5901 hCoV-19/USA/TX-QDX-1199/2020\|EPI_ISL_572037\|2020-0 | .................... | .................... | .................... | .................... | .................... | .................... | |
| L | 5902 hCoV-19/USA/TX-QDX-1201/2020\|EPI_ISL_572038\|2020-0 | .................... | .................... | .................... | .................... | .................... | .................... | |
| L | 5903 hCoV-19/USA/TX-QDX-1202/2020\|EPI_ISL_572039\|2020-0 | .................... | .................... | .................... | .................... | .................... | .................... | |
| L | 5904 hCoV-19/USA/TX-QDX-1285/2020\|EPI_ISL_572040\|2020-0 | .................... | .................... | .................... | .................... | .................... | .................... | |
| L | 5905 hCoV-19/USA/UT-QDX-1275/2020\|EPI_ISL_572041\|2020-0 | .................... | .................... | .................... | .................... | .................... | .................... | |
| L | 5906 hCoV-19/USA/UT-QDX-1276/2020\|EPI_ISL_572042\|2020-0 | .................... | .................... | .................... | .................... | .................... | .................... | |
| L | 5907 hCoV-19/USA/UT-QDX-1277/2020\|EPI_ISL_572043\|2020-0 | .................... | .................... | .................... | .................... | .................... | .................... | |
| L | 5908 hCoV-19/USA/UT-QDX-1278/2020\|EPI_ISL_572044\|2020-0 | .................... | .................... | .................... | .................... | .................... | .................... | |
| L | 5909 hCoV-19/USA/UT-QDX-1279/2020\|EPI_ISL_572045\|2020-0 | .................... | .................... | .................... | .................... | .................... | .................... | |
| L | 5910 hCoV-19/USA/UT-QDX-1280/2020\|EPI_ISL_572046\|2020-0 | .................... | .................... | .................... | .................... | .................... | .................... | |
| L | 5911 hCoV-19/USA/UT-QDX-1281/2020\|EPI_ISL_572047\|2020-0 | .................... | .................... | .................... | .................... | .................... | .................... | |
| L | 5912 hCoV-19/USA/UT-QDX-1282/2020\|EPI_ISL_572048\|2020-0 | .................... | .................... | .................... | .................... | .................... | .................... | |
| L | 5913 hCoV-19/USA/UT-QDX-1283/2020\|EPI_ISL_572049\|2020-0 | .................... | .................... | .................... | .................... | .................... | .................... | |
| L | 5914 hCoV-19/USA/WA-QDX-1270/2020\|EPI_ISL_572050\|2020-0 | .................... | .................... | .................... | .................... | .................... | .................... | |
| L | 5915 hCoV-19/USA/WA-QDX-1271/2020\|EPI_ISL_572051\|2020-0 | .................... | .................... | .................... | .................... | .................... | .................... | |
| L | 5916 hCoV-19/USA/WA-QDX-1273/2020\|EPI_ISL_572052\|2020-0 | .................... | .................... | .................... | .................... | .................... | .................... | |
| L | 5917 hCoV-19/USA/WV-QDX-1284/2020\|EPI_ISL_572053\|2020-0 | .................... | .................... | .................... | .................... | .................... | .................... | |
| L | 5918 hCoV-19/USA/AR-QDX-1315/2020\|EPI_ISL_572054\|2020-0 | .................... | .................... | .................... | .................... | .................... | .................... | |
| L | 5919 hCoV-19/USA/CA-QDX-1395/2020\|EPI_ISL_572055\|2020-0 | .................... | .................... | .................... | .................... | .................... | .................... | |
| L | 5920 hCoV-19/USA/CA-QDX-1396/2020\|EPI_ISL_572056\|2020-0 | .................... | .................... | .................... | .................... | .................... | .................... | |
| L | 5921 hCoV-19/USA/CA-QDX-1397/2020\|EPI_ISL_572057\|2020-0 | .................... | .................... | .................... | .................... | .................... | .................... | |
| L | 5922 hCoV-19/USA/CA-QDX-1398/2020\|EPI_ISL_572058\|2020-0 | .................... | .................... | .................... | .................... | .................... | .................... | |
| L | 5923 hCoV-19/USA/CA-QDX-1400/2020\|EPI_ISL_572059\|2020-0 | .................... | .................... | .................... | .................... | .................... | .................... | |
| L | 5924 hCoV-19/USA/CA-QDX-1408/2020\|EPI_ISL_572060\|2020-0 | .................... | .................... | .................... | .................... | .................... | .................... | |
| L | 5925 hCoV-19/USA/CA-QDX-1409/2020\|EPI_ISL_572061\|2020-0 | .................... | .................... | .................... | .................... | .................... | .................... | |
| L | 5926 hCoV-19/USA/CA-QDX-1410/2020\|EPI_ISL_572062\|2020-0 | .................... | .................... | .................... | .................... | .................... | .................... | |
| L | 5927 hCoV-19/USA/CA-QDX-1411/2020\|EPI_ISL_572063\|2020-0 | .................... | .................... | .................... | .................... | .................... | .................... | |
| L | 5928 hCoV-19/USA/CA-QDX-1412/2020\|EPI_ISL_572064\|2020-0 | .................... | .................... | .................... | .................... | .................... | .................... | |
| L | 5929 hCoV-19/USA/CA-QDX-1413/2020\|EPI_ISL_572065\|2020-0 | .................... | .................... | .................... | .................... | .................... | .................... | |
| L | 5930 hCoV-19/USA/CA-QDX-1414/2020\|EPI_ISL_572066\|2020-0 | .................... | .................... | .................... | .................... | .................... | .................... | |
| L | 5931 hCoV-19/USA/CA-QDX-1415/2020\|EPI_ISL_572067\|2020-0 | .................... | .................... | .................... | .................... | .................... | .................... | |
| L | 5932 hCoV-19/USA/CA-QDX-1416/2020\|EPI_ISL_572068\|2020-0 | .................... | .................... | .................... | .................... | .................... | .................... | |
| L | 5933 hCoV-19/USA/CA-QDX-1417/2020\|EPI_ISL_572069\|2020-0 | .................... | .................... | .................... | .................... | .................... | .................... | |
| L | 5934 hCoV-19/USA/CA-QDX-1418/2020\|EPI_ISL_572070\|2020-0 | .................... | .................... | .................... | .................... | .................... | .................... | |
| L | 5935 hCoV-19/USA/CA-QDX-1419/2020\|EPI_ISL_572071\|2020-0 | .................... | .................... | .................... | .................... | .................... | .................... | |
| L | 5936 hCoV-19/USA/CA-QDX-1420/2020\|EPI_ISL_572072\|2020-0 | .................... | .................... | .................... | .................... | .................... | .................... | |
| L | 5937 hCoV-19/USA/CA-QDX-1422/2020\|EPI_ISL_572074\|2020-0 | .................... | .................... | .................... | .................... | .................... | .................... | |
| L | 5938 hCoV-19/USA/CA-QDX-1423/2020\|EPI_ISL_572075\|2020-0 | .................... | .................... | .................... | .................... | .................... | .................... | |
| L | 5939 hCoV-19/USA/CA-QDX-1424/2020\|EPI_ISL_572076\|2020-0 | .................... | .................... | .................... | .................... | .................... | .................... | |
| L | 5940 hCoV-19/USA/CA-QDX-1425/2020\|EPI_ISL_572077\|2020-0 | .................... | .................... | .................... | .................... | .................... | .................... | |
| L | 5941 hCoV-19/USA/CA-QDX-1426/2020\|EPI_ISL_572078\|2020-0 | .................... | .................... | .................... | .................... | .................... | .................... | |
| L | 5942 hCoV-19/USA/CA-QDX-1427/2020\|EPI_ISL_572079\|2020-0 | .................... | .................... | .................... | .................... | .................... | .................... | |
| L | 5943 hCoV-19/USA/CA-QDX-1428/2020\|EPI_ISL_572080\|2020-0 | .................... | .................... | .................... | .................... | .................... | .................... | |
| L | 5944 hCoV-19/USA/CA-QDX-1429/2020\|EPI_ISL_572081\|2020-0 | .................... | .................... | .................... | .................... | .................... | .................... | |
| L | 5945 hCoV-19/USA/CA-QDX-1430/2020\|EPI_ISL_572082\|2020-0 | .................... | .................... | .................... | .................... | .................... | .................... | |
| L | 5946 hCoV-19/USA/CA-QDX-1431/2020\|EPI_ISL_572083\|2020-0 | .................... | .................... | .................... | .................... | .................... | .................... | |
| L | 5947 hCoV-19/USA/CA-QDX-1432/2020\|EPI_ISL_572084\|2020-0 | .................... | .................... | .................... | .................... | .................... | .................... | |
| L | 5948 hCoV-19/USA/CA-QDX-1433/2020\|EPI_ISL_572085\|2020-0 | .................... | .................... | .................... | .................... | .................... | .................... | |
| L | 5949 hCoV-19/USA/CA-QDX-1434/2020\|EPI_ISL_572086\|2020-0 | .................... | .................... | .................... | .................... | .................... | .................... | |
| L | 5950 hCoV-19/USA/CA-QDX-1435/2020\|EPI_ISL_572087\|2020-0 | .................... | .................... | .................... | .................... | .................... | .................... | |
| L | 5951 hCoV-19/USA/CA-QDX-1436/2020\|EPI_ISL_572088\|2020-0 | .................... | .................... | .................... | .................... | .................... | .................... | |
| L | 5952 hCoV-19/USA/CA-QDX-1437/2020\|EPI_ISL_572089\|2020-0 | .................... | .................... | .................... | .................... | .................... | .................... | |
| L | 5953 hCoV-19/USA/CA-QDX-1438/2020\|EPI_ISL_572090\|2020-0 | .................... | .................... | .................... | .................... | .................... | .................... | |
| L | 5954 hCoV-19/USA/CA-QDX-1440/2020\|EPI_ISL_572091\|2020-0 | .................... | .................... | .................... | .................... | .................... | .................... | |
| L | 5955 hCoV-19/USA/CA-QDX-1441/2020\|EPI_ISL_572092\|2020-0 | .................... | .................... | .................... | .................... | .................... | .................... | |
| L | 5956 hCoV-19/USA/CA-QDX-1442/2020\|EPI_ISL_572093\|2020-0 | .................... | .................... | .................... | .................... | .................... | .................... | |
| L | 5957 hCoV-19/USA/CA-QDX-1443/2020\|EPI_ISL_572094\|2020-0 | .................... | .................... | .................... | .................... | .................... | .................... | |
| L | 5958 hCoV-19/USA/CA-QDX-1444/2020\|EPI_ISL_572095\|2020-0 | .................... | .................... | .................... | .................... | .................... | .................... | |
| L | 5959 hCoV-19/USA/CA-QDX-1445/2020\|EPI_ISL_572096\|2020-0 | .................... | .................... | .................... | .................... | .................... | .................... | |
| L | 5960 hCoV-19/USA/CT-QDX-1401/2020\|EPI_ISL_572097\|2020-0 | .................... | .................... | .................... | .................... | .................... | .................... | |
| L | 5961 hCoV-19/USA/CT-QDX-1402/2020\|EPI_ISL_572098\|2020-0 | .................... | .................... | .................... | .................... | .................... | .................... | |

| | | | | | | | |
|---|---|---|---|---|---|---|---|
| L | 5962 hCoV-19/USA/CT-QDX-1403/2020\|EPI_ISL_572099\|2020-0 | ···················· | ···················· | ····················· | ···················· | ···················· | ···················· |
| L | 5964 hCoV-19/England/MILK-9C737C/2020\|EPI_ISL_573301\|20 | ···················· | ···················· | ····················· | ···················· | ···················· | ···················· |
| L | 5965 hCoV-19/England/MILK-9C738B/2020\|EPI_ISL_573302\|20 | ···················· | ···················· | ····················· | ···················· | ···················· | ···················· |
| L | 5966 hCoV-19/England/MILK-9C739A/2020\|EPI_ISL_573303\|20 | ···················· | ···················· | ····················· | ···················· | ···················· | ···················· |
| L | 5967 hCoV-19/England/MILK-9C743D/2020\|EPI_ISL_573304\|20 | ···················· | ···················· | ····················· | ···················· | ···················· | ···················· |
| L | 5968 hCoV-19/England/MILK-9C744C/2020\|EPI_ISL_573305\|20 | ···················· | ···················· | ····················· | ···················· | ···················· | ···················· |
| L | 5969 hCoV-19/England/MILK-9C745B/2020\|EPI_ISL_573306\|20 | ···················· | ···················· | ····················· | ···················· | ···················· | ···················· |
| L | 5970 hCoV-19/England/MILK-9C746A/2020\|EPI_ISL_573307\|20 | ···················· | ···················· | ····················· | ···················· | ···················· | ···················· |
| L | 5971 hCoV-19/England/MILK-9C7479/2020\|EPI_ISL_573308\|20 | ···················· | ···················· | ····················· | ···················· | ···················· | ···················· |
| L | 5972 hCoV-19/England/MILK-9C74A6/2020\|EPI_ISL_573309\|20 | ···················· | ·A·················· | ····················· | ···················· | ···················· | ···················· |
| L | 5973 hCoV-19/England/MILK-9C75FE/2020\|EPI_ISL_573310\|20 | ···················· | ···················· | ····················· | ···················· | ···················· | ···················· |
| L | 5974 hCoV-19/England/NORT-2947FB/2020\|EPI_ISL_573355\|20 | ···················· | ···················· | ····················· | ···················· | ···················· | ···················· |
| L | 5975 hCoV-19/England/NORT-294816/2020\|EPI_ISL_573356\|20 | ···················· | ···················· | ····················· | ···················· | ···················· | ···················· |
| L | 5976 hCoV-19/England/NORT-29C3E0/2020\|EPI_ISL_573357\|20 | ···················· | ···················· | ····················· | ···················· | ···················· | ···················· |
| L | 5977 hCoV-19/England/NORT-2A09D8/2020\|EPI_ISL_573358\|20 | ···················· | ···················· | ····················· | ···················· | ···················· | ···················· |
| L | 5978 hCoV-19/England/NORT-2A0A11/2020\|EPI_ISL_573359\|20 | ···················· | ···················· | ····················· | ···················· | ···················· | ···················· |
| L | 5979 hCoV-19/England/NORT-2A0AD5/2020\|EPI_ISL_573360\|20 | ···················· | ···················· | ····················· | ···················· | ···················· | ···················· |
| L | 5980 hCoV-19/England/NORT-2A0BF0/2020\|EPI_ISL_573361\|20 | ···················· | ···················· | ····················· | ···················· | ···················· | ···················· |
| L | 5982 hCoV-19/England/NORT-2A0CA2/2020\|EPI_ISL_573363\|20 | ···················· | ···················· | ····················· | ···················· | ···················· | ···················· |
| L | 5984 hCoV-19/England/NORT-2A0D18/2020\|EPI_ISL_573365\|20 | ···················· | ···················· | ····················· | ···················· | ···················· | ···················· |
| L | 5986 hCoV-19/England/NORT-2A0D63/2020\|EPI_ISL_573367\|20 | ···················· | ···················· | ····················· | ···················· | ···················· | ···················· |
| L | 5987 hCoV-19/England/NORT-2A0D72/2020\|EPI_ISL_573368\|20 | ···················· | ···················· | ····················· | ···················· | ···················· | ···················· |
| L | 5988 hCoV-19/England/NORT-2A0E06/2020\|EPI_ISL_573369\|20 | ···················· | ···················· | ····················· | ···················· | ···················· | ···················· |
| L | 5989 hCoV-19/England/NORT-2A0E15/2020\|EPI_ISL_573370\|20 | ···················· | ···················· | ····················· | ···················· | ···················· | ···················· |
| L | 5990 hCoV-19/England/NORT-2A0E24/2020\|EPI_ISL_573371\|20 | ···················· | ···················· | ····················· | ···················· | ···················· | ···················· |
| L | 5991 hCoV-19/England/NORT-2A4330/2020\|EPI_ISL_573372\|20 | ···················· | ···················· | ····················· | ···················· | ···················· | ···················· |
| L | 5992 hCoV-19/England/NORT-2A438B/2020\|EPI_ISL_573373\|20 | ···················· | ·A·················· | ····················· | ···················· | ···················· | ···················· |
| L | 5994 hCoV-19/England/NORT-2A450D/2020\|EPI_ISL_573375\|20 | ···················· | ···················· | ····················· | ···················· | ···················· | ···················· |
| L | 5995 hCoV-19/England/NORT-2A45C1/2020\|EPI_ISL_573376\|20 | ···················· | ···················· | ····················· | ···················· | ····T··············· | ···················· |
| L | 5996 hCoV-19/England/NORT-2A4682/2020\|EPI_ISL_573377\|20 | ···················· | ···················· | ····················· | ···················· | ···················· | ···················· |
| L | 5997 hCoV-19/England/NORT-2A4691/2020\|EPI_ISL_573378\|20 | ···················· | ···················· | ····················· | ···················· | ···················· | ···················· |
| L | 5998 hCoV-19/England/NORT-2A46DD/2020\|EPI_ISL_573379\|20 | ···················· | ···················· | ····················· | ···················· | ···················· | ···················· |
| L | 5999 hCoV-19/England/NORT-2A46FB/2020\|EPI_ISL_573380\|20 | ···················· | ···················· | ····················· | ···················· | ···················· | ···················· |
| L | 6000 hCoV-19/England/NORT-2A4716/2020\|EPI_ISL_573381\|20 | ···················· | ···················· | ····················· | ···················· | ····T··············· | ···················· |
| L | 6001 hCoV-19/England/NORT-2A4804/2020\|EPI_ISL_573382\|20 | ···················· | ···················· | ····················· | ···················· | ···················· | ···················· |
| L | 6002 hCoV-19/England/NORT-2A4831/2020\|EPI_ISL_573383\|20 | ···················· | ···················· | ····················· | ···················· | ····T··············· | ···················· |
| L | 6003 hCoV-19/England/NORT-2A4840/2020\|EPI_ISL_573384\|20 | ···················· | ···················· | ····················· | ···················· | ···················· | ···················· |
| L | 6004 hCoV-19/England/NORT-2A485F/2020\|EPI_ISL_573385\|20 | ···················· | ···················· | ····················· | ···················· | ···················· | ···················· |
| L | 6005 hCoV-19/England/NORT-2A488C/2020\|EPI_ISL_573386\|20 | ···················· | ···················· | ····················· | ···················· | ····T··············· | ···················· |
| L | 6006 hCoV-19/England/NORT-2A489B/2020\|EPI_ISL_573387\|20 | ···················· | ···················· | ····················· | ···················· | ····T··············· | ···················· |
| L | 6007 hCoV-19/England/NORT-2A48F5/2020\|EPI_ISL_573388\|20 | ···················· | ···················· | ····················· | ···················· | ···················· | ···················· |
| L | 6008 hCoV-19/England/NORT-2A493E/2020\|EPI_ISL_573389\|20 | ···················· | ···················· | ····················· | ···················· | ···················· | ···················· |
| L | 6009 hCoV-19/England/NORT-2A494D/2020\|EPI_ISL_573390\|20 | ···················· | ···················· | ····················· | ···················· | ···················· | ···················· |
| L | 6010 hCoV-19/England/NORT-2A497A/2020\|EPI_ISL_573391\|20 | ···················· | ···················· | ····················· | ···················· | ····T··············· | ···················· |
| L | 6011 hCoV-19/England/NORT-2A49E3/2020\|EPI_ISL_573392\|20 | ···················· | ···················· | ····················· | ···················· | ···················· | ···················· |
| L | 6012 hCoV-19/England/NORT-2A4A1D/2020\|EPI_ISL_573393\|20 | ···················· | ···················· | ····················· | ···················· | ···················· | ···················· |
| L | 6013 hCoV-19/England/NORT-2A4AC2/2020\|EPI_ISL_573394\|20 | ···················· | ···················· | ····················· | ···················· | ···················· | ···················· |
| L | 6014 hCoV-19/England/NORT-2A4AD1/2020\|EPI_ISL_573395\|20 | ···················· | ···················· | ····················· | ···················· | ···················· | ···················· |
| L | 6015 hCoV-19/England/NORT-2A4B1A/2020\|EPI_ISL_573396\|20 | ···················· | ···················· | ····················· | ···················· | ···················· | ···················· |
| L | 6016 hCoV-19/England/NORT-2A4B65/2020\|EPI_ISL_573397\|20 | ···················· | ···················· | ····················· | ···················· | ···················· | ···················· |
| L | 6017 hCoV-19/England/NORT-2A4B83/2020\|EPI_ISL_573398\|20 | ···················· | ···················· | ····················· | ···················· | ····T··············· | ···················· |
| L | 6018 hCoV-19/England/NORT-2A4B92/2020\|EPI_ISL_573399\|20 | ···················· | ···················· | ····················· | ···················· | ···················· | ···················· |
| L | 6019 hCoV-19/Indonesia/PA-EIJK09/2020\|EPI_ISL_574603\|20 | ···················· | ···················· | ····················· | ···················· | ···················· | ···················· |
| L | 6020 hCoV-19/Indonesia/SA-EIJK13/2020\|EPI_ISL_574604\|20 | ···················· | ···················· | ····················· | ···················· | ···················· | ···················· |
| L | 6021 hCoV-19/Indonesia/YO-EIJK15/2020\|EPI_ISL_574605\|20 | ···················· | ···················· | ····················· | ···················· | ···················· | ···················· |
| L | 6022 hCoV-19/Indonesia/JB-EIJK16/2020\|EPI_ISL_574606\|20 | ···················· | ···················· | ····················· | ···················· | ···················· | ···················· |
| L | 6023 hCoV-19/Indonesia/JK-EIJK19/2020\|EPI_ISL_574607\|20 | ···················· | ···················· | ····················· | ···················· | ···················· | ···················· |
| L | 6024 hCoV-19/Indonesia/JK-EIJK20/2020\|EPI_ISL_574608\|20 | ···················· | ···················· | ····················· | ···················· | ···················· | ···················· |
| L | 6025 hCoV-19/Indonesia/JK-EIJK24/2020\|EPI_ISL_574609\|20 | ···················· | ···················· | ····················· | ···················· | ···················· | ···················· |
| L | 6026 hCoV-19/Indonesia/JK-EIJK34/2020\|EPI_ISL_574610\|20 | ···················· | ···················· | ····················· | ···················· | ···················· | ···················· |
| L | 6027 hCoV-19/Indonesia/BT-EIJK36/2020\|EPI_ISL_574611\|20 | ···················· | ···················· | ····················· | ···················· | ···················· | ···················· |
| L | 6028 hCoV-19/Indonesia/JB-EIJK41/2020\|EPI_ISL_574612\|20 | ···················· | ···················· | ····················· | ···················· | ···················· | ···················· |
| L | 6029 hCoV-19/Indonesia/JK-EIJK42/2020\|EPI_ISL_574613\|20 | ···················· | ···················· | ····················· | ···················· | ···················· | ···················· |
| L | 6030 hCoV-19/Indonesia/JK-EIJK45/2020\|EPI_ISL_574614\|20 | ···················· | ···················· | ····················· | ···················· | ···················· | ···················· |
| L | 6031 hCoV-19/Indonesia/JK-EIJK46/2020\|EPI_ISL_574615\|20 | ···················· | ···················· | ····················· | ···················· | ···················· | ···················· |
| L | 6032 hCoV-19/Indonesia/JK-EIJK47/2020\|EPI_ISL_574616\|20 | ···················· | ···················· | ····················· | ···················· | ···················· | ···················· |
| L | 6033 hCoV-19/Indonesia/JK-EIJK48/2020\|EPI_ISL_574617\|20 | ···················· | ···················· | ····················· | ···················· | ···················· | ···················· |
| L | 6034 hCoV-19/Indonesia/JK-EIJK49/2020\|EPI_ISL_574618\|20 | ···················· | ····A··············· | ····················· | ···················· | ···················· | ···················· |
| L | 6035 hCoV-19/Indonesia/SA-EIJK50/2020\|EPI_ISL_574619\|20 | ···················· | ···················· | ····················· | ···················· | ···················· | ···················· |
| L | 6036 hCoV-19/Indonesia/SA-EIJK51/2020\|EPI_ISL_574620\|20 | ···················· | ···················· | ····················· | ···················· | ···················· | ···················· |
| L | 6037 hCoV-19/Indonesia/SA-EIJK52/2020\|EPI_ISL_574621\|20 | ···················· | ···················· | ····················· | ···················· | ···················· | ···················· |
| L | 6038 hCoV-19/Indonesia/SA-EIJK53/2020\|EPI_ISL_574622\|20 | ···················· | ···················· | ····················· | ···················· | ···················· | ···················· |
| L | 6039 hCoV-19/Indonesia/SA-EIJK54/2020\|EPI_ISL_574623\|20 | ···················· | ···················· | ····················· | ···················· | ···················· | ···················· |
| L | 6040 hCoV-19/USA/WA-S2844/2020\|EPI_ISL_574686\|2020-09-1 | ···················· | ·············A······ | ····················· | ···················· | ···················· | ···················· |
| L | 6041 hCoV-19/USA/WA-S2845/2020\|EPI_ISL_574687\|2020-09-1 | ···················· | ···················· | ····················· | ···················· | ···················· | ···················· |
| L | 6042 hCoV-19/USA/WA-S2846/2020\|EPI_ISL_574688\|2020-09-1 | ···················· | ···················· | ····················· | ···················· | ···················· | ···················· |
| L | 6043 hCoV-19/USA/WA-S2847/2020\|EPI_ISL_574689\|2020-09-1 | ···················· | ···················· | ····················· | ···················· | ···················· | ···················· |
| L | 6044 hCoV-19/USA/WA-S2848/2020\|EPI_ISL_574690\|2020-09-1 | ···················· | ···················· | ····················· | ···················· | ···················· | ···················· |
| L | 6045 hCoV-19/USA/WA-S2849/2020\|EPI_ISL_574691\|2020-09-1 | ···················· | ···················· | ····················· | ···················· | ···················· | ···················· |
| L | 6046 hCoV-19/England/204020544/2020\|EPI_ISL_574697\|2020 | ···················· | ·A·················· | ····················· | ···················· | ···················· | ···A················ |

| | | | | | | | |
|---|---|---|---|---|---|---|---|
| L | 6047 hCoV-19/England/204020545/2020\|EPI_ISL_574698\|2020 | ···················· | ·A·················· | ····················· | ···················· | ···················· | ··A················· |
| L | 6048 hCoV-19/England/204020546/2020\|EPI_ISL_574699\|2020 | ···················· | ·A·················· | ····················· | ···················· | ···················· | ··A················· |
| L | 6049 hCoV-19/England/MILK-9763E5/2020\|EPI_ISL_575930\|20 | ···················· | ···················· | ····················· | ···················· | ···················· | ···················· |
| L | 6050 hCoV-19/England/MILK-9733AC/2020\|EPI_ISL_575932\|20 | ···················· | ···················· | ····················· | ···················· | ···················· | ···················· |
| L | 6051 hCoV-19/England/MILK-97650D/2020\|EPI_ISL_575934\|20 | ···················· | ·A·················· | ····················· | ···················· | ···················· | ···················· |
| L | 6052 hCoV-19/England/MILK-9735F1/2020\|EPI_ISL_575935\|20 | ···················· | ···················· | ····················· | ···················· | ···················· | ···················· |
| L | 6053 hCoV-19/England/MILK-9730A5/2020\|EPI_ISL_575936\|20 | ···················· | ···A················ | ····················· | ···················· | ···················· | ···················· |
| L | 6054 hCoV-19/England/MILK-97639A/2020\|EPI_ISL_575937\|20 | ···················· | ···················· | ····················· | ···················· | ···················· | ···················· |
| L | 6055 hCoV-19/England/MILK-97462A/2020\|EPI_ISL_575938\|20 | ···················· | ···················· | ····················· | ···················· | ···················· | ···················· |
| L | 6056 hCoV-19/England/MILK-973263/2020\|EPI_ISL_575939\|20 | ···················· | ···················· | ····················· | ···················· | ···················· | ···················· |
| L | 6057 hCoV-19/England/MILK-9764C4/2020\|EPI_ISL_575940\|20 | ···················· | ···················· | ····················· | ···················· | ···················· | ···················· |
| L | 6058 hCoV-19/England/MILK-974833/2020\|EPI_ISL_575941\|20 | ···················· | C··················· | ····················· | ···················· | ···················· | ···················· |
| L | 6059 hCoV-19/Wales/MILK-97312A/2020\|EPI_ISL_575942\|2020 | ···················· | ···················· | ····················· | ···················· | ···················· | ···················· |
| L | 6060 hCoV-19/England/MILK-974BC1/2020\|EPI_ISL_575943\|20 | ···················· | ·A·················· | ····················· | ···················· | ···················· | ···················· |
| L | 6061 hCoV-19/England/MILK-9731FD/2020\|EPI_ISL_575944\|20 | ···················· | ···················· | ····················· | ···················· | ···················· | ···················· |
| L | 6062 hCoV-19/England/MILK-974BD0/2020\|EPI_ISL_575945\|20 | ···················· | ···················· | ····················· | ···················· | ···················· | ···················· |
| L | 6063 hCoV-19/England/MILK-974772/2020\|EPI_ISL_575946\|20 | ···················· | ···················· | ····················· | ···················· | ···················· | ···················· |
| L | 6064 hCoV-19/England/MILK-97637C/2020\|EPI_ISL_575947\|20 | ···················· | ···················· | ····················· | ···················· | ···················· | ···················· |
| L | 6065 hCoV-19/England/MILK-9746EE/2020\|EPI_ISL_575948\|20 | ···················· | ···················· | ····················· | ···················· | ···················· | ···················· |
| L | 6066 hCoV-19/England/MILK-973658/2020\|EPI_ISL_575949\|20 | ···················· | ···················· | ····················· | ···················· | ···················· | ···················· |
| L | 6067 hCoV-19/England/MILK-97660A/2020\|EPI_ISL_575950\|20 | ···················· | ···················· | ····················· | ···················· | ···················· | ···················· |
| L | 6068 hCoV-19/England/MILK-97641F/2020\|EPI_ISL_575951\|20 | ···················· | ···················· | ····················· | ···················· | ···················· | ···················· |
| L | 6069 hCoV-19/England/MILK-97497C/2020\|EPI_ISL_575952\|20 | ···················· | ·A·················· | ····················· | ···················· | ···················· | ··A················· |
| L | 6070 hCoV-19/England/MILK-97487F/2020\|EPI_ISL_575953\|20 | ···················· | ···················· | ····················· | ···················· | ···················· | ···················· |
| L | 6071 hCoV-19/England/MILK-9767F8/2020\|EPI_ISL_575954\|20 | ···················· | ·R·················· | ····················· | ···················· | ···················· | ···················· |
| L | 6072 hCoV-19/England/MILK-9747DC/2020\|EPI_ISL_575959\|20 | ···················· | ···················· | ····················· | ···················· | ···················· | ···················· |
| L | 6073 hCoV-19/England/MILK-97495E/2020\|EPI_ISL_575961\|20 | ···················· | ···················· | ····················· | ···················· | ···················· | ···················· |
| L | 6074 hCoV-19/England/MILK-9768D7/2020\|EPI_ISL_575962\|20 | ···················· | ···················· | ····················· | ···················· | ···················· | ···················· |
| L | 6075 hCoV-19/England/MILK-974921/2020\|EPI_ISL_575963\|20 | ···················· | ···················· | ····················· | ···················· | ···················· | ···················· |
| L | 6076 hCoV-19/England/MILK-9765C1/2020\|EPI_ISL_575964\|20 | ···················· | ···················· | ····················· | ···················· | ···················· | ···················· |
| L | 6077 hCoV-19/England/MILK-9730F0/2020\|EPI_ISL_575965\|20 | ···················· | ···················· | ····················· | ···················· | ···················· | ···················· |
| L | 6078 hCoV-19/England/MILK-974745/2020\|EPI_ISL_575967\|20 | ···················· | ···················· | ····················· | ···················· | ···················· | ···················· |
| L | 6079 hCoV-19/England/MILK-974815/2020\|EPI_ISL_575968\|20 | ···················· | ···················· | ····················· | ···················· | ···················· | ···················· |
| L | 6080 hCoV-19/England/MILK-97496D/2020\|EPI_ISL_575970\|20 | ···················· | ···················· | ····················· | ···················· | ···················· | ···················· |
| L | 6081 hCoV-19/England/MILK-9767DA/2020\|EPI_ISL_575971\|20 | ···················· | ···················· | ····················· | ···················· | ···················· | ···················· |
| L | 6082 hCoV-19/England/MILK-9767AD/2020\|EPI_ISL_575972\|20 | ···················· | ···················· | ····················· | ···················· | ···················· | ···················· |
| L | 6083 hCoV-19/USA/MI-UM-10033781516/2020\|EPI_ISL_578107\| | ···················· | ···················· | ····················· | ···················· | ···················· | ···················· |
| L | 6084 hCoV-19/USA/MI-UM-10033781702/2020\|EPI_ISL_578108\| | ···················· | ···················· | ····················· | ···················· | ···················· | ···················· |
| L | 6085 hCoV-19/USA/MI-UM-10033782000/2020\|EPI_ISL_578109\| | ···················· | ···················· | ····················· | ···················· | ···················· | ···················· |
| L | 6086 hCoV-19/USA/MI-UM-10033783681/2020\|EPI_ISL_578110\| | ···················· | ···················· | ····················· | ···················· | ···················· | ···················· |
| L | 6088 hCoV-19/USA/MI-UM-10033785805/2020\|EPI_ISL_578112\| | ···················· | ···················· | ····················· | ···················· | ···················· | ···················· |
| L | 6090 hCoV-19/USA/MI-UM-10033792025/2020\|EPI_ISL_578114\| | ···················· | ···················· | ····················· | ···················· | ···················· | ···················· |
| L | 6091 hCoV-19/USA/MI-UM-10033792343/2020\|EPI_ISL_578115\| | ···················· | ···················· | ····················· | ···················· | ···················· | ···················· |
| L | 6092 hCoV-19/USA/MI-UM-10033792567/2020\|EPI_ISL_578116\| | ···················· | ···················· | ····················· | ···················· | ···················· | ···················· |
| L | 6094 hCoV-19/USA/MI-UM-10033793198/2020\|EPI_ISL_578118\| | ···················· | ···················· | ····················· | ···················· | ···················· | ···················· |
| L | 6097 hCoV-19/USA/MI-UM-10033808445/2020\|EPI_ISL_578124\| | ···················· | ···················· | ····················· | ···················· | ···················· | ···················· |
| L | 6100 hCoV-19/New Zealand/20VR3149/2020\|EPI_ISL_579440\|2 | ···················· | ···················· | ····················· | ···················· | ···················· | ···················· |
| L | 6101 hCoV-19/New Zealand/20VR3150/2020\|EPI_ISL_579441\|2 | ···················· | ···················· | ····················· | ···················· | ···················· | ···················· |
| L | 6104 hCoV-19/New Zealand/20VR3154/2020\|EPI_ISL_579444\|2 | ···················· | ···················· | ····················· | ···················· | ···················· | ···················· |
| L | 6105 hCoV-19/New Zealand/20VR3155/2020\|EPI_ISL_579445\|2 | ···················· | ···················· | ····················· | ···················· | ···················· | ···················· |
| L | 6108 hCoV-19/New Zealand/20VR3158/2020\|EPI_ISL_579448\|2 | ···················· | ···················· | ····················· | ···················· | ···················· | ···················· |
| L | 6111 hCoV-19/New Zealand/20VR3164/2020\|EPI_ISL_579454\|2 | ···················· | ···················· | ····················· | ···················· | ···················· | ···················· |
| L | 6112 hCoV-19/England/ALDP-9E6DC4/2020\|EPI_ISL_580403\|20 | ···················· | ···················· | ····················· | ···················· | ···················· | ···················· |
| L | 6113 hCoV-19/England/ALDP-9E6B60/2020\|EPI_ISL_580404\|20 | ···················· | ···················· | ····················· | ···················· | ···················· | ···················· |
| L | 6114 hCoV-19/England/ALDP-9E72E2/2020\|EPI_ISL_580406\|20 | ···················· | ···················· | ····················· | ···················· | ···················· | ···················· |
| L | 6115 hCoV-19/England/ALDP-9E709D/2020\|EPI_ISL_580407\|20 | ···················· | ···················· | ····················· | ···················· | ···················· | ···················· |
| L | 6116 hCoV-19/England/ALDP-9E60F8/2020\|EPI_ISL_580408\|20 | ···················· | ···················· | ····················· | ···················· | ···················· | ···················· |
| L | 6117 hCoV-19/England/ALDP-9E682D/2020\|EPI_ISL_580410\|20 | ···················· | ···················· | ····················· | ···················· | ···················· | ···················· |
| L | 6118 hCoV-19/England/ALDP-9E66F6/2020\|EPI_ISL_580411\|20 | ···················· | ···················· | ····················· | ···················· | ···················· | ···················· |
| L | 6119 hCoV-19/England/ALDP-9E6CB8/2020\|EPI_ISL_580412\|20 | ···················· | ···················· | ····················· | ···················· | ···················· | ···················· |
| L | 6120 hCoV-19/England/ALDP-9E73A3/2020\|EPI_ISL_580413\|20 | ···················· | ···················· | ····················· | ···················· | ···················· | ···················· |
| L | 6121 hCoV-19/England/CAMC-9EC60F/2020\|EPI_ISL_580414\|20 | ···················· | ···················· | ····················· | ···················· | ···················· | ···················· |
| L | 6122 hCoV-19/England/ALDP-9E61D7/2020\|EPI_ISL_580415\|20 | ···················· | ···················· | ····················· | ···················· | ···················· | ···················· |
| L | 6123 hCoV-19/England/ALDP-9E7042/2020\|EPI_ISL_580416\|20 | ·········T·········· | ···················· | ····················· | ···················· | ···················· | ···················· |
| L | 6124 hCoV-19/England/ALDP-9E62A7/2020\|EPI_ISL_580417\|20 | ···················· | ···················· | ····················· | ···················· | ···················· | ···················· |
| L | 6125 hCoV-19/England/CAMC-9EC292/2020\|EPI_ISL_580418\|20 | ···················· | ·A·················· | ····················· | ···················· | ···················· | ··A················· |
| L | 6126 hCoV-19/England/ALDP-9E69A2/2020\|EPI_ISL_580419\|20 | ···················· | ···················· | ····················· | ···················· | ···················· | ···················· |
| L | 6127 hCoV-19/Switzerland/BL-42205193/2020\|EPI_ISL_58173 | ···················· | ···················· | ····················· | ···················· | ···················· | ···················· |
| L | 6128 hCoV-19/Switzerland/BL-42205226/2020\|EPI_ISL_58173 | ···················· | ···················· | ····················· | ···················· | ···················· | ···················· |
| L | 6130 hCoV-19/Switzerland/BS-42205161/2020\|EPI_ISL_58173 | ···················· | ···················· | ····················· | ···················· | ···················· | ···················· |
| L | 6132 hCoV-19/Switzerland/BS-42206107/2020\|EPI_ISL_58174 | ···················· | ···················· | ····················· | ···················· | ···················· | ···················· |
| L | 6133 hCoV-19/Switzerland/BS-42206243/2020\|EPI_ISL_58174 | ···················· | ···················· | ····················· | ···················· | ···················· | ···················· |
| L | 6134 hCoV-19/Switzerland/BS-42206244/2020\|EPI_ISL_58174 | ···················· | ···················· | ····················· | ···················· | ···················· | ···················· |
| L | 6135 hCoV-19/Switzerland/BS-42206307/2020\|EPI_ISL_58174 | ···················· | ···················· | ····················· | ···················· | ···················· | ···················· |
| L | 6136 hCoV-19/Switzerland/BS-42206310/2020\|EPI_ISL_58174 | ···················· | ···················· | ····················· | ···················· | ···················· | ···················· |
| L | 6138 hCoV-19/Switzerland/BS-42209636/2020\|EPI_ISL_58180 | ···················· | ···················· | ····················· | ···················· | ···················· | ···················· |
| L | 6139 hCoV-19/Switzerland/BL-42209645/2020\|EPI_ISL_58180 | ···················· | ···················· | ····················· | ···················· | ···················· | ···················· |
| L | 6142 hCoV-19/Switzerland/BS-42210895/2020\|EPI_ISL_58180 | ···················· | ···················· | ····················· | ···················· | ···················· | ···················· |
| L | 6143 hCoV-19/Switzerland/BS-42210896/2020\|EPI_ISL_58180 | ···················· | ···················· | ····················· | ···················· | ···················· | ···················· |
| L | 6145 hCoV-19/USA/NY-QDX-1224/2020\|EPI_ISL_572000\|2020-0 | ···················· | ···················· | ····················· | ···················· | ···················· | ···················· |

| | | | | | | | | |
|---|---|---|---|---|---|---|---|---|
| L | 6146 | hCoV-19/USA/NY-QDX-1225/2020\|EPI_ISL_572001\|2020-0 | ···················· | ···················· | ···················· | ···················· | ···················· | ···················· |
| L | 6147 | hCoV-19/USA/NY-QDX-1226/2020\|EPI_ISL_572002\|2020-0 | ···················· | ···················· | ···················· | ···················· | ···················· | ···················· |
| L | 6148 | hCoV-19/USA/NY-QDX-1227/2020\|EPI_ISL_572003\|2020-0 | ···················· | ···················· | ···················· | ···················· | ···················· | ···················· |
| L | 6149 | hCoV-19/USA/NY-QDX-1228/2020\|EPI_ISL_572004\|2020-0 | ···················· | ···················· | ···················· | ···················· | ···················· | ···················· |
| L | 6150 | hCoV-19/USA/NY-QDX-1229/2020\|EPI_ISL_572005\|2020-0 | ···················· | ···················· | ···················· | ···················· | ···················· | ···················· |
| L | 6151 | hCoV-19/USA/NY-QDX-1230/2020\|EPI_ISL_572006\|2020-0 | ···················· | ···················· | ···················· | ···················· | ···················· | ···················· |
| L | 6152 | hCoV-19/USA/NY-QDX-1231/2020\|EPI_ISL_572007\|2020-0 | ···················· | ···················· | ···················· | ···················· | ···················· | ···················· |
| L | 6153 | hCoV-19/USA/NY-QDX-1232/2020\|EPI_ISL_572008\|2020-0 | ···················· | ···················· | ···················· | ···················· | ···················· | ···················· |
| L | 6154 | hCoV-19/USA/NY-QDX-1233/2020\|EPI_ISL_572009\|2020-0 | ···················· | ···················· | ···················· | ···················· | ···················· | ···················· |
| L | 6155 | hCoV-19/USA/NY-QDX-1234/2020\|EPI_ISL_572010\|2020-0 | ···················· | ···················· | ···················· | ···················· | ···················· | ···················· |
| L | 6156 | hCoV-19/USA/NY-QDX-1235/2020\|EPI_ISL_572011\|2020-0 | ···················· | ···················· | ···················· | ···················· | ···················· | ···················· |
| L | 6157 | hCoV-19/USA/NY-QDX-1236/2020\|EPI_ISL_572012\|2020-0 | ···················· | ···················· | ···················· | ···················· | ···················· | ···················· |
| L | 6158 | hCoV-19/USA/NY-QDX-1237/2020\|EPI_ISL_572013\|2020-0 | ···················· | ···················· | ···················· | ···················· | ···················· | ···················· |
| L | 6159 | hCoV-19/USA/NY-QDX-1238/2020\|EPI_ISL_572014\|2020-0 | ···················· | ···················· | ···················· | ···················· | ···················· | ···················· |
| L | 6160 | hCoV-19/USA/NY-QDX-1239/2020\|EPI_ISL_572015\|2020-0 | ···················· | ···················· | ···················· | ···················· | ···················· | ···················· |
| L | 6161 | hCoV-19/USA/NY-QDX-1240/2020\|EPI_ISL_572016\|2020-0 | ···················· | ···················· | ···················· | ···················· | ···················· | ···················· |
| L | 6162 | hCoV-19/USA/NY-QDX-1250/2020\|EPI_ISL_572017\|2020-0 | ···················· | ···················· | ···················· | ···················· | ···················· | ···················· |
| L | 6163 | hCoV-19/USA/NY-QDX-1251/2020\|EPI_ISL_572018\|2020-0 | ···················· | ···················· | ···················· | ···················· | ···················· | ···················· |
| L | 6164 | hCoV-19/USA/NY-QDX-1255/2020\|EPI_ISL_572019\|2020-0 | ···················· | ···················· | ···················· | ···················· | ···················· | ···················· |
| L | 6165 | hCoV-19/USA/NY-QDX-1256/2020\|EPI_ISL_572020\|2020-0 | ···················· | ···················· | ···················· | ···················· | ···················· | ···················· |
| L | 6166 | hCoV-19/USA/NY-QDX-1257/2020\|EPI_ISL_572021\|2020-0 | ···················· | ···················· | ···················· | ···················· | ···················· | ···················· |
| L | 6167 | hCoV-19/USA/NY-QDX-1258/2020\|EPI_ISL_572022\|2020-0 | ···················· | ···················· | ···················· | ···················· | ···················· | ···················· |
| L | 6168 | hCoV-19/USA/NY-QDX-1259/2020\|EPI_ISL_572023\|2020-0 | ···················· | ···················· | ···················· | ···················· | ···················· | ···················· |
| L | 6169 | hCoV-19/England/NORT-1B641AF/2020\|EPI_ISL_573311\|2 | ···················· | ···················· | ···················· | ···················· | ···················· | ···················· |
| L | 6170 | hCoV-19/England/NORT-286149/2020\|EPI_ISL_573312\|20 | ···················· | ···················· | ···················· | ···················· | ···················· | ···················· |
| L | 6171 | hCoV-19/England/NORT-286167/2020\|EPI_ISL_573313\|20 | ···················· | ···················· | ···················· | ···················· | ···················· | ···················· |
| L | 6172 | hCoV-19/England/NORT-286176/2020\|EPI_ISL_573314\|20 | ···················· | ···················· | ···················· | ···················· | ···················· | ···················· |
| L | 6173 | hCoV-19/England/NORT-286185/2020\|EPI_ISL_573315\|20 | ···················· | ···················· | ···················· | ···················· | ···················· | ···················· |
| L | 6174 | hCoV-19/England/NORT-2861A3/2020\|EPI_ISL_573316\|20 | ···················· | ···················· | ···················· | ···················· | ···················· | ···················· |
| L | 6175 | hCoV-19/England/NORT-2861D0/2020\|EPI_ISL_573317\|20 | ···················· | ···················· | ···················· | ···················· | ···················· | ···················· |
| L | 6176 | hCoV-19/England/NORT-2861EF/2020\|EPI_ISL_573318\|20 | ···················· | ···················· | ···················· | ···················· | ···················· | ···················· |
| L | 6177 | hCoV-19/England/NORT-2861FE/2020\|EPI_ISL_573319\|20 | ···················· | ···················· | ···················· | ···················· | ···················· | ···················· |
| L | 6178 | hCoV-19/England/NORT-286219/2020\|EPI_ISL_573320\|20 | ···················· | ···················· | ···················· | ···················· | ···················· | ···················· |
| L | 6179 | hCoV-19/England/NORT-286273/2020\|EPI_ISL_573321\|20 | ···················· | ···················· | ···················· | ···················· | ···················· | ···················· |
| L | 6180 | hCoV-19/England/NORT-286282/2020\|EPI_ISL_573322\|20 | ···················· | ···················· | ···················· | ···················· | ···················· | ···················· |
| L | 6181 | hCoV-19/England/NORT-28CBDC/2020\|EPI_ISL_573323\|20 | ···················· | ···················· | ···················· | ···················· | ···················· | ···················· |
| L | 6183 | hCoV-19/England/NORT-293D09/2020\|EPI_ISL_573325\|20 | ···················· | ···················· | ···················· | ···················· | ···················· | ···················· |
| L | 6184 | hCoV-19/England/NORT-293D45/2020\|EPI_ISL_573326\|20 | ···················· | ···················· | ···················· | ···················· | ···················· | ···················· |
| L | 6185 | hCoV-19/England/NORT-293D54/2020\|EPI_ISL_573327\|20 | ···················· | ···················· | ···················· | ···················· | ···················· | ···················· |
| L | 6186 | hCoV-19/England/NORT-293DDC/2020\|EPI_ISL_573328\|20 | ···················· | ···················· | ···················· | ···················· | ···················· | ···················· |
| L | 6187 | hCoV-19/England/NORT-293E06/2020\|EPI_ISL_573329\|20 | ···················· | ···················· | ···················· | ···················· | ···················· | ···················· |
| L | 6188 | hCoV-19/England/NORT-293E24/2020\|EPI_ISL_573330\|20 | ···················· | ···················· | ···················· | ···················· | ···················· | ···················· |
| L | 6189 | hCoV-19/England/NORT-293E60/2020\|EPI_ISL_573331\|20 | ···················· | ···················· | ···················· | ···················· | ···················· | ···················· |
| L | 6191 | hCoV-19/England/NORT-293FB8/2020\|EPI_ISL_573333\|20 | ···················· | ···················· | ···················· | ···················· | ···················· | ···················· |
| L | 6192 | hCoV-19/England/NORT-293FF4/2020\|EPI_ISL_573334\|20 | ···················· | ···················· | ···················· | ···················· | ···················· | ···················· |
| L | 6193 | hCoV-19/England/NORT-29400F/2020\|EPI_ISL_573335\|20 | ···················· | ···················· | ···················· | ···················· | ···················· | ···················· |
| L | 6194 | hCoV-19/England/NORT-29412A/2020\|EPI_ISL_573336\|20 | ···················· | ···················· | ···················· | ···················· | ···················· | ···················· |
| L | 6197 | hCoV-19/England/NORT-294193/2020\|EPI_ISL_573339\|20 | ···················· | ···················· | ···················· | ···················· | ···················· | ···················· |
| L | 6198 | hCoV-19/England/NORT-2941A2/2020\|EPI_ISL_573340\|20 | ···················· | ···················· | ···················· | ···················· | ···················· | ···················· |
| L | 6199 | hCoV-19/England/NORT-294254/2020\|EPI_ISL_573341\|20 | ···················· | ···················· | ···················· | ···················· | ···················· | ···················· |
| L | 6200 | hCoV-19/England/NORT-2943D9/2020\|EPI_ISL_573342\|20 | ···················· | ···················· | ···················· | ···················· | ···················· | ···················· |
| L | 6201 | hCoV-19/England/NORT-294421/2020\|EPI_ISL_573343\|20 | ···················· | ···················· | ···················· | ···················· | ···················· | ···················· |
| L | 6202 | hCoV-19/England/NORT-29447C/2020\|EPI_ISL_573344\|20 | ···················· | ···················· | ···················· | ···················· | ···················· | ···················· |
| L | 6203 | hCoV-19/England/NORT-29448B/2020\|EPI_ISL_573345\|20 | ···················· | ···················· | ···················· | ···················· | ···················· | ···················· |
| L | 6204 | hCoV-19/England/NORT-294579/2020\|EPI_ISL_573346\|20 | ···················· | ···················· | ···················· | ···················· | ···················· | ···················· |
| L | 6205 | hCoV-19/England/NORT-2945A6/2020\|EPI_ISL_573347\|20 | ···················· | ···················· | ···················· | ···················· | ···················· | ···················· |
| L | 6206 | hCoV-19/England/NORT-2945C4/2020\|EPI_ISL_573348\|20 | ···················· | ···················· | ···················· | ···················· | ···················· | ···················· |
| L | 6207 | hCoV-19/England/NORT-29463A/2020\|EPI_ISL_573349\|20 | ···················· | ···················· | ···················· | ···················· | ···················· | ···················· |
| L | 6208 | hCoV-19/England/NORT-294667/2020\|EPI_ISL_573350\|20 | ···················· | ···················· | ···················· | ···················· | ···················· | ···················· |
| L | 6209 | hCoV-19/England/NORT-294676/2020\|EPI_ISL_573351\|20 | ···················· | ···················· | ···················· | ···················· | ···················· | ···················· |
| L | 6210 | hCoV-19/England/NORT-294694/2020\|EPI_ISL_573352\|20 | ···················· | ···················· | ···················· | ···················· | ···················· | ···················· |
| L | 6211 | hCoV-19/England/NORT-294728/2020\|EPI_ISL_573353\|20 | ···················· | ···················· | ···················· | ···················· | ···················· | ···················· |
| L | 6212 | hCoV-19/England/NORT-294737/2020\|EPI_ISL_573354\|20 | ···················· | ···················· | ···················· | ···················· | ···················· | ···················· |
| L | 6213 | hCoV-19/USA/WA-S2806/2020\|EPI_ISL_574648\|2020-05-1 | ···················· | ···················· | ···················· | ···················· | ···················· | ···················· |
| L | 6214 | hCoV-19/USA/WA-S2807/2020\|EPI_ISL_574649\|2020-08-2 | ···················· | ···················· | ···················· | ···················· | ···················· | ···················· |
| L | 6215 | hCoV-19/USA/WA-S2808/2020\|EPI_ISL_574650\|2020-08-3 | ···················· | ·A·················· | ···················· | ···················· | ···················· | ···················· |
| L | 6216 | hCoV-19/USA/WA-S2810/2020\|EPI_ISL_574652\|2020-09-0 | ···················· | ···················· | ···················· | ···················· | ···················· | ···················· |
| L | 6217 | hCoV-19/USA/WA-S2811/2020\|EPI_ISL_574653\|2020-09-0 | ···················· | ·A·················· | ···················· | ···················· | ···················· | ···················· |
| L | 6218 | hCoV-19/USA/WA-S2812/2020\|EPI_ISL_574654\|2020-09-0 | ···················· | ·A·················· | ···················· | ···················· | ···················· | ···················· |
| L | 6219 | hCoV-19/USA/WA-S2813/2020\|EPI_ISL_574655\|2020-09-0 | ···················· | ·A·················· | ···················· | ···················· | ···················· | ···················· |
| L | 6220 | hCoV-19/USA/WA-S2815/2020\|EPI_ISL_574657\|2020-09-0 | ···················· | ···················· | ···················· | ···················· | ···················· | ···················· |
| L | 6221 | hCoV-19/USA/WA-S2816/2020\|EPI_ISL_574658\|2020-09-0 | ···················· | ·A·················· | ···················· | ···················· | ···················· | ···················· |
| L | 6222 | hCoV-19/USA/WA-S2818/2020\|EPI_ISL_574660\|2020-09-0 | ···················· | ···················· | ···················· | ···················· | ···················· | ···················· |
| L | 6223 | hCoV-19/USA/WA-S2819/2020\|EPI_ISL_574661\|2020-09-0 | ···················· | ·A·················· | ···················· | ···················· | ···················· | ···················· |
| L | 6224 | hCoV-19/USA/WA-S2820/2020\|EPI_ISL_574662\|2020-09-0 | ···················· | ···················· | ···················· | ···················· | ···················· | ···················· |
| L | 6225 | hCoV-19/USA/WA-S2821/2020\|EPI_ISL_574663\|2020-09-0 | ···················· | ·A·················· | ···················· | ···················· | ···················· | ···················· |
| L | 6226 | hCoV-19/USA/WA-S2822/2020\|EPI_ISL_574664\|2020-09-0 | ···················· | ·A·················· | ···················· | ···················· | ···················· | ···················· |
| L | 6227 | hCoV-19/USA/WA-S2823/2020\|EPI_ISL_574665\|2020-09-0 | ···················· | ·A·················· | ···················· | ···················· | ···················· | ···················· |
| L | 6228 | hCoV-19/USA/WA-S2824/2020\|EPI_ISL_574666\|2020-09-0 | ···················· | ·A·················· | ···················· | ···················· | ···················· | ···················· |
| L | 6229 | hCoV-19/USA/WA-S2825/2020\|EPI_ISL_574667\|2020-09-0 | ···················· | ·A·················· | ···················· | ···················· | ···················· | ···················· |

| | | | | | | | | |
|---|---|---|---|---|---|---|---|---|
| L | 6230 | hCoV-19/USA/WA-S2826/2020\|EPI_ISL_574668\|2020-09-0 | ···················· | ···················· | ···················· | ···················· | ···················· | ···················· |
| L | 6231 | hCoV-19/USA/WA-S2827/2020\|EPI_ISL_574669\|2020-09-0 | ···················· | ···················· | ···················· | ···················· | ···················· | ···················· |
| L | 6232 | hCoV-19/USA/WA-S2828/2020\|EPI_ISL_574670\|2020-09-0 | ···················· | ···················· | ···················· | ···················· | ···················· | ···················· |
| L | 6233 | hCoV-19/USA/WA-S2830/2020\|EPI_ISL_574672\|2020-09-0 | ···················· | ···················· | ···················· | ···················· | ···················· | ···················· |
| L | 6234 | hCoV-19/USA/WA-S2831/2020\|EPI_ISL_574673\|2020-09-0 | ···················· | ···················· | ···················· | ···················· | ···················· | ···················· |
| L | 6235 | hCoV-19/USA/WA-S2832/2020\|EPI_ISL_574674\|2020-09-0 | ···················· | ···················· | ···················· | ···················· | ···················· | ···················· |
| L | 6236 | hCoV-19/USA/WA-S2833/2020\|EPI_ISL_574675\|2020-09-0 | ···················· | ···················· | ···················· | ···················· | ···················· | ···················· |
| L | 6237 | hCoV-19/USA/WA-S2834/2020\|EPI_ISL_574676\|2020-09-1 | ···················· | ·A·················· | ···················· | ···················· | ···················· | ···················· |
| L | 6238 | hCoV-19/USA/WA-S2835/2020\|EPI_ISL_574677\|2020-09-1 | ···················· | ···················· | ···················· | ···················· | ···················· | ···················· |
| L | 6239 | hCoV-19/USA/WA-S2836/2020\|EPI_ISL_574678\|2020-09-1 | ···················· | ···················· | ···················· | ···················· | ···················· | ···················· |
| L | 6240 | hCoV-19/USA/WA-S2837/2020\|EPI_ISL_574679\|2020-09-1 | ···················· | ·A·················· | ···················· | ···················· | ···················· | ···················· |
| L | 6241 | hCoV-19/USA/WA-S2838/2020\|EPI_ISL_574680\|2020-09-1 | ···················· | ·············A······ | ···················· | ···················· | ···················· | ···················· |
| L | 6242 | hCoV-19/USA/WA-S2839/2020\|EPI_ISL_574681\|2020-09-1 | ···················· | ···················· | ···················· | ···················· | ···················· | ···················· |
| L | 6243 | hCoV-19/USA/WA-S2840/2020\|EPI_ISL_574682\|2020-09-1 | ···················· | ·A·················· | ···················· | ···················· | ···················· | ···················· |
| L | 6244 | hCoV-19/USA/WA-S2842/2020\|EPI_ISL_574684\|2020-09-1 | ···················· | ·A·················· | ···················· | ···················· | ···················· | ···················· |
| L | 6245 | hCoV-19/USA/WA-S2843/2020\|EPI_ISL_574685\|2020-09-1 | ···················· | ·············A······ | ···················· | ···················· | ···················· | ···················· |
| L | 6246 | hCoV-19/England/MILK-974657/2020\|EPI_ISL_575955\|20 | ···················· | ···················· | ···················· | ···················· | ···················· | ···················· |
| L | 6247 | hCoV-19/England/MILK-973254/2020\|EPI_ISL_575956\|20 | ···················· | ···················· | ···················· | ···················· | ···················· | ···················· |
| L | 6248 | hCoV-19/England/MILK-97353D/2020\|EPI_ISL_575958\|20 | ···················· | ···················· | ···················· | ···················· | ···················· | ···················· |
| L | 6249 | hCoV-19/England/MILK-97652B/2020\|EPI_ISL_575973\|20 | ···················· | ···················· | ···················· | ···················· | ···················· | ···················· |
| L | 6250 | hCoV-19/England/MILK-97361C/2020\|EPI_ISL_575975\|20 | ···················· | ···················· | ···················· | ···················· | ···················· | ···················· |
| L | 6251 | hCoV-19/England/MILK-9764D3/2020\|EPI_ISL_575976\|20 | ···················· | ···················· | ···················· | ···················· | ···················· | ···················· |
| L | 6252 | hCoV-19/England/MILK-973166/2020\|EPI_ISL_575977\|20 | ···················· | ···················· | ···················· | ···················· | ···················· | ···················· |
| L | 6253 | hCoV-19/England/MILK-97356A/2020\|EPI_ISL_575978\|20 | ···················· | ···················· | ···················· | ···················· | ···················· | ···················· |
| L | 6254 | hCoV-19/England/MILK-976260/2020\|EPI_ISL_575980\|20 | ···················· | ···················· | ···················· | ···················· | ···················· | ···················· |
| L | 6255 | hCoV-19/England/MILK-9735D3/2020\|EPI_ISL_575981\|20 | ···················· | ···················· | ···················· | ···················· | ···················· | ···················· |
| L | 6256 | hCoV-19/Scotland/ALDP-9CD25B/2020\|EPI_ISL_575982\|2 | ···················· | ···················· | ···················· | ···················· | ···················· | ···················· |
| L | 6257 | hCoV-19/England/ALDP-9CDC8A/2020\|EPI_ISL_575983\|20 | ···················· | ·A·················· | ···················· | ···················· | ···················· | ···················· |
| L | 6258 | hCoV-19/Scotland/MILK-9C263C/2020\|EPI_ISL_575984\|2 | ···················· | ···················· | ···················· | ···················· | ···················· | ···················· |
| L | 6259 | hCoV-19/Scotland/MILK-9C261E/2020\|EPI_ISL_575985\|2 | ···················· | ···················· | ···················· | ···················· | ·····T·············· | ···················· |
| L | 6260 | hCoV-19/Scotland/MILK-9C412A/2020\|EPI_ISL_575986\|2 | ···················· | ···················· | ···················· | ···················· | ···················· | ···················· |
| L | 6261 | hCoV-19/Scotland/MILK-9C3A4E/2020\|EPI_ISL_575987\|2 | ···················· | ···················· | ···················· | ···················· | ···················· | ···················· |
| L | 6262 | hCoV-19/Scotland/MILK-9C2F31/2020\|EPI_ISL_575988\|2 | ···················· | ···················· | ···················· | ···················· | ···················· | ···················· |
| L | 6263 | hCoV-19/Scotland/MILK-9C41FD/2020\|EPI_ISL_575989\|2 | ···················· | ·A·················· | ···················· | ···················· | ···················· | ··A················· |
| L | 6264 | hCoV-19/Scotland/MILK-9C3817/2020\|EPI_ISL_575990\|2 | ···················· | ···················· | ···················· | ···················· | ···················· | ···················· |
| L | 6265 | hCoV-19/Scotland/MILK-9C3783/2020\|EPI_ISL_575991\|2 | ···················· | ···················· | ···················· | ···················· | ···················· | ···················· |
| L | 6266 | hCoV-19/Scotland/MILK-9C30D3/2020\|EPI_ISL_575992\|2 | ···················· | ···················· | ···················· | ···················· | ···················· | ···················· |
| L | 6267 | hCoV-19/Scotland/MILK-9C4218/2020\|EPI_ISL_575993\|2 | ···················· | ·A·················· | ···················· | ···················· | ···················· | ··A················· |
| L | 6268 | hCoV-19/Scotland/QEUH-9C7CCC/2020\|EPI_ISL_575996\|2 | ···················· | ···················· | ···················· | ···················· | ···················· | ···················· |
| L | 6269 | hCoV-19/Scotland/QEUH-9C9091/2020\|EPI_ISL_575997\|2 | ···················· | ···················· | ···················· | ···················· | ···················· | ···················· |
| L | 6270 | hCoV-19/Scotland/QEUH-9C7D50/2020\|EPI_ISL_575999\|2 | ···················· | ···················· | ···················· | ···················· | ···················· | ···················· |
| L | 6271 | hCoV-19/Netherlands/ZH-EMC-580/2020\|EPI_ISL_578008 | ···················· | ···A················ | ···················· | ···················· | ···················· | ···················· |
| L | 6272 | hCoV-19/Netherlands/ZH-EMC-581/2020\|EPI_ISL_578009 | ···················· | ···················· | ···················· | ···················· | ·····T·············· | ···················· |
| L | 6273 | hCoV-19/Netherlands/ZH-EMC-583/2020\|EPI_ISL_578011 | ···················· | ···················· | ···················· | ···················· | ·····T·············· | ···················· |
| L | 6274 | hCoV-19/Netherlands/ZH-EMC-585/2020\|EPI_ISL_578013 | ···················· | ···················· | ···················· | ···················· | ···················· | ···················· |
| L | 6275 | hCoV-19/Netherlands/ZH-EMC-586/2020\|EPI_ISL_578014 | ···················· | ···················· | ···················· | ···················· | ···················· | ···················· |
| L | 6276 | hCoV-19/Netherlands/ZH-EMC-587/2020\|EPI_ISL_578015 | ···················· | ···················· | ···················· | ···················· | ·····T·············· | ···················· |
| L | 6277 | hCoV-19/Netherlands/ZH-EMC-588/2020\|EPI_ISL_578016 | ···················· | ···················· | ···················· | ···················· | ···················· | ···················· |
| L | 6278 | hCoV-19/Netherlands/ZH-EMC-589/2020\|EPI_ISL_578017 | ···················· | ···················· | ···················· | ···················· | ···················· | ···················· |
| L | 6279 | hCoV-19/Netherlands/ZH-EMC-590/2020\|EPI_ISL_578018 | ···················· | ···················· | ···················· | ···················· | ···················· | ···················· |
| L | 6280 | hCoV-19/Netherlands/ZH-EMC-591/2020\|EPI_ISL_578019 | ···················· | ···················· | ···················· | ···················· | ···················· | ···················· |
| L | 6281 | hCoV-19/Netherlands/ZH-EMC-592/2020\|EPI_ISL_578020 | ···················· | ···················· | ···················· | ···················· | ···················· | ···················· |
| L | 6282 | hCoV-19/Netherlands/ZH-EMC-593/2020\|EPI_ISL_578021 | ···················· | ···················· | ···················· | ···················· | ···················· | ···················· |
| L | 6283 | hCoV-19/Netherlands/ZH-EMC-594/2020\|EPI_ISL_578022 | ···················· | ···················· | ···················· | ···················· | ···················· | ···················· |
| L | 6284 | hCoV-19/Netherlands/ZH-EMC-595/2020\|EPI_ISL_578023 | ···················· | ···················· | ···················· | ···················· | ···················· | ···················· |
| L | 6285 | hCoV-19/Netherlands/ZH-EMC-596/2020\|EPI_ISL_578024 | ···················· | ···················· | ···················· | ···················· | ···················· | ···················· |
| L | 6286 | hCoV-19/Netherlands/ZH-EMC-597/2020\|EPI_ISL_578025 | ···················· | ···················· | ···················· | ···················· | ···················· | ···················· |
| L | 6287 | hCoV-19/Netherlands/ZH-EMC-598/2020\|EPI_ISL_578026 | ···················· | ···················· | ···················· | ···················· | ···················· | ···················· |
| L | 6288 | hCoV-19/Netherlands/ZH-EMC-599/2020\|EPI_ISL_578027 | ···················· | ···················· | ···················· | ···················· | ···················· | ···················· |
| L | 6289 | hCoV-19/Netherlands/ZH-EMC-600/2020\|EPI_ISL_578028 | ···················· | ···················· | ···················· | ···················· | ···················· | ···················· |
| L | 6290 | hCoV-19/Netherlands/ZH-EMC-601/2020\|EPI_ISL_578029 | ···················· | ···················· | ···················· | ···················· | ·····T·············· | ···················· |
| L | 6291 | hCoV-19/Netherlands/ZH-EMC-605/2020\|EPI_ISL_578033 | ···················· | ···················· | ···················· | ···················· | ···················· | ···················· |
| L | 6292 | hCoV-19/Netherlands/ZH-EMC-606/2020\|EPI_ISL_578034 | ···················· | ···················· | ···················· | ···················· | ···················· | ···················· |
| L | 6293 | hCoV-19/Netherlands/ZH-EMC-609/2020\|EPI_ISL_578037 | ···················· | ···················· | ···················· | ···················· | ···················· | ···················· |
| L | 6294 | hCoV-19/Netherlands/ZH-EMC-610/2020\|EPI_ISL_578038 | ···················· | ···················· | ···················· | ···················· | ·····T·············· | ···················· |
| L | 6295 | hCoV-19/Netherlands/ZH-EMC-611/2020\|EPI_ISL_578039 | ···················· | ···················· | ···················· | ···················· | ···················· | ···················· |
| L | 6296 | hCoV-19/Netherlands/ZH-EMC-612/2020\|EPI_ISL_578040 | ···················· | ···················· | ···················· | ···················· | ···················· | ···················· |
| L | 6297 | hCoV-19/Netherlands/ZH-EMC-613/2020\|EPI_ISL_578041 | ···················· | ···················· | ···················· | ···················· | ·····T·············· | ···················· |
| L | 6298 | hCoV-19/Netherlands/ZH-EMC-614/2020\|EPI_ISL_578042 | ···················· | ···················· | ···················· | ···················· | ···················· | ···················· |
| L | 6299 | hCoV-19/Netherlands/ZH-EMC-615/2020\|EPI_ISL_578043 | ···················· | ···················· | ···················· | ···················· | ···················· | ···················· |
| L | 6300 | hCoV-19/Netherlands/ZH-EMC-616/2020\|EPI_ISL_578044 | ···················· | ···················· | ···················· | ···················· | ···················· | ···················· |
| L | 6301 | hCoV-19/Netherlands/ZH-EMC-617/2020\|EPI_ISL_578045 | ···················· | ···················· | ···················· | ···················· | ···················· | ···················· |
| L | 6302 | hCoV-19/Netherlands/ZH-EMC-618/2020\|EPI_ISL_578046 | ···················· | ···A················ | ···················· | ···················· | ···················· | ···················· |
| L | 6303 | hCoV-19/Netherlands/ZH-EMC-619/2020\|EPI_ISL_578047 | ···················· | ···················· | ···················· | ···················· | ···················· | ···················· |
| L | 6304 | hCoV-19/Netherlands/ZH-EMC-620/2020\|EPI_ISL_578048 | ···················· | ···················· | ···················· | ···················· | ···················· | ···················· |
| L | 6306 | hCoV-19/New Zealand/20VR2723/2020\|EPI_ISL_579341\|2 | ···················· | ···················· | ···················· | ···················· | ···················· | ···················· |
| L | 6310 | hCoV-19/New Zealand/20VR2918/2020\|EPI_ISL_579349\|2 | ···················· | ···················· | ···················· | ···················· | ···················· | ···················· |
| L | 6311 | hCoV-19/New Zealand/20VR2919/2020\|EPI_ISL_579350\|2 | ···················· | ···················· | ···················· | ···················· | ···················· | ···················· |
| L | 6315 | hCoV-19/New Zealand/20VR2933/2020\|EPI_ISL_579357\|2 | ···················· | ···················· | ···················· | ···················· | ···················· | ···················· |
| L | 6316 | hCoV-19/New Zealand/20VR2934/2020\|EPI_ISL_579358\|2 | ···················· | ···················· | ···················· | ···················· | ···················· | ···················· |

| | | | | | | | | |
|---|---|---|---|---|---|---|---|---|
| L | 6318 hCoV-19/New Zealand/20VR2943/2020\|EPI_ISL_579364\|2 | ···················· | ···················· | ···················· | ···················· | ···················· | ···················· | |
| L | 6319 hCoV-19/New Zealand/20VR2944/2020\|EPI_ISL_579365\|2 | ···················· | ···················· | ···················· | ···················· | ···················· | ···················· | |
| L | 6320 hCoV-19/New Zealand/20VR2955/2020\|EPI_ISL_579374\|2 | ···················· | ···················· | ···················· | ···················· | ···················· | ···················· | |
| L | 6323 hCoV-19/New Zealand/20VR2959/2020\|EPI_ISL_579378\|2 | ···················· | ···················· | ···················· | ···················· | ···················· | ···················· | |
| L | 6324 hCoV-19/New Zealand/20VR3007/2020\|EPI_ISL_579379\|2 | ···················· | ···················· | ···················· | ···················· | ···················· | ···················· | |
| L | 6325 hCoV-19/England/ALDP-9E66C9/2020\|EPI_ISL_580320\|20 | ···················· | ···················· | ···················· | ···················· | ···················· | ···················· | |
| L | 6326 hCoV-19/England/ALDP-9E669C/2020\|EPI_ISL_580321\|20 | ···················· | ···················· | ···················· | ···················· | ···················· | ···················· | |
| L | 6327 hCoV-19/England/CAMC-9EC1C2/2020\|EPI_ISL_580322\|20 | ···················· | ···················· | ···················· | ···················· | ···················· | ···················· | |
| L | 6328 hCoV-19/England/ALDP-9E717C/2020\|EPI_ISL_580323\|20 | ···················· | ···················· | ···················· | ···················· | ···················· | ···················· | |
| L | 6329 hCoV-19/England/ALDP-9E715E/2020\|EPI_ISL_580324\|20 | ···················· | ···················· | ···················· | ···················· | ···················· | ···················· | |
| L | 6330 hCoV-19/England/CAMC-9EC03E/2020\|EPI_ISL_580325\|20 | ···················· | ···················· | ···················· | ···················· | ···················· | ···················· | |
| L | 6331 hCoV-19/England/ALDP-9E6CD6/2020\|EPI_ISL_580326\|20 | ···················· | ···················· | ···················· | ···················· | ···················· | ···················· | |
| L | 6332 hCoV-19/England/CAMC-9EC696/2020\|EPI_ISL_580327\|20 | ···················· | ···················· | ···················· | ···················· | ···················· | ···················· | |
| L | 6333 hCoV-19/England/ALDP-9E72F1/2020\|EPI_ISL_580330\|20 | ···················· | ···················· | ···················· | ···················· | ···················· | ···················· | |
| L | 6334 hCoV-19/England/ALDP-9E725B/2020\|EPI_ISL_580331\|20 | ···················· | ···················· | ···················· | ···················· | ···················· | ···················· | |
| L | 6335 hCoV-19/England/ALDP-9E6E3A/2020\|EPI_ISL_580334\|20 | ···················· | ···················· | ···················· | ···················· | ···················· | ···················· | |
| L | 6336 hCoV-19/England/ALDP-9E716D/2020\|EPI_ISL_580335\|20 | ···················· | ···················· | ···················· | ···················· | ···················· | ···················· | |
| L | 6337 hCoV-19/England/CAMC-9EC63C/2020\|EPI_ISL_580340\|20 | ···················· | ···················· | ···················· | ···················· | ···················· | ···················· | |
| L | 6338 hCoV-19/England/CAMC-9EC5C6/2020\|EPI_ISL_580341\|20 | ···················· | ·A·················· | ···················· | ···················· | ···················· | ···A················ | |
| L | 6339 hCoV-19/England/ALDP-9E60AD/2020\|EPI_ISL_580342\|20 | ···················· | ···················· | ···················· | ···················· | ···················· | ···················· | |
| L | 6340 hCoV-19/England/ALDP-9E6A09/2020\|EPI_ISL_580343\|20 | ···················· | ···················· | ···················· | ···················· | ···················· | ···················· | |
| L | 6341 hCoV-19/England/ALDP-9E7419/2020\|EPI_ISL_580345\|20 | ···················· | ···················· | ···················· | ···················· | ···················· | ···················· | |
| L | 6342 hCoV-19/England/ALDP-9E6D3D/2020\|EPI_ISL_580346\|20 | ···················· | ···················· | ···················· | ···················· | ···················· | ···················· | |
| L | 6343 hCoV-19/England/ALDP-9E6ED0/2020\|EPI_ISL_580347\|20 | ···················· | ···················· | ···················· | ···················· | ···················· | ···················· | |
| L | 6344 hCoV-19/England/ALDP-9E6869/2020\|EPI_ISL_580348\|20 | ···················· | ···················· | ···················· | ···················· | ···················· | ···················· | |
| L | 6345 hCoV-19/England/ALDP-9E673F/2020\|EPI_ISL_580349\|20 | ···················· | ···················· | ···················· | ···················· | ···················· | ···················· | |
| L | 6346 hCoV-19/England/ALDP-9E6D79/2020\|EPI_ISL_580350\|20 | ···················· | ···················· | ···················· | ···················· | ···················· | ···················· | |
| L | 6347 hCoV-19/England/ALDP-9E6B7F/2020\|EPI_ISL_580351\|20 | ···················· | ···················· | ···················· | ···················· | ···················· | ···················· | |
| L | 6348 hCoV-19/England/ALDP-9E7006/2020\|EPI_ISL_580352\|20 | ···················· | ···················· | ···················· | ···················· | ···················· | ···················· | |
| L | 6349 hCoV-19/Wales/CAMC-9EC089/2020\|EPI_ISL_580353\|2020 | ···················· | ···················· | ···················· | ···················· | ···················· | ···················· | |
| L | 6350 hCoV-19/England/CAMC-9EC71B/2020\|EPI_ISL_580354\|20 | ···················· | ···················· | ···················· | ···················· | ···················· | ···················· | |
| L | 6351 hCoV-19/England/ALDP-9E6720/2020\|EPI_ISL_580355\|20 | ···················· | ···················· | ···················· | ···················· | ···················· | ···················· | |
| L | 6352 hCoV-19/England/ALDP-9E73D0/2020\|EPI_ISL_580356\|20 | ···················· | ···················· | ···················· | ···················· | ···················· | ···················· | |
| L | 6353 hCoV-19/England/ALDP-9E6FBF/2020\|EPI_ISL_580357\|20 | ···················· | ···················· | ···················· | ···················· | ···················· | ···················· | |
| L | 6354 hCoV-19/England/ALDP-9E67D5/2020\|EPI_ISL_580358\|20 | ···················· | ···················· | ···················· | ···················· | ···················· | ···················· | |
| L | 6355 hCoV-19/England/ALDP-9E6A45/2020\|EPI_ISL_580359\|20 | ···················· | ···················· | ···················· | ···················· | ···················· | ···················· | |
| L | 6356 hCoV-19/England/ALDP-9E70AC/2020\|EPI_ISL_580360\|20 | ···················· | ···················· | ···················· | ···················· | ···················· | ···················· | |
| L | 6357 hCoV-19/England/ALDP-9E6A27/2020\|EPI_ISL_580361\|20 | ···················· | ···················· | ···················· | ···················· | ···················· | ···················· | |
| L | 6358 hCoV-19/Belgium/ULG-10486/2020\|EPI_ISL_581650\|2020 | ···················· | ···················· | ···················· | ···················· | ···················· | ···················· | |
| L | 6361 hCoV-19/Belgium/ULG-10490/2020\|EPI_ISL_581653\|2020 | ···················· | ···················· | ···················· | ···················· | ···················· | ···················· | |
| L | 6362 hCoV-19/Belgium/ULG-10492/2020\|EPI_ISL_581654\|2020 | ···················· | ···················· | ···················· | ···················· | ···················· | ···················· | |
| L | 6363 hCoV-19/Belgium/ULG-10493/2020\|EPI_ISL_581655\|2020 | ···················· | ···················· | ···················· | ···················· | ···················· | ···················· | |
| L | 6367 hCoV-19/Belgium/ULG-10497/2020\|EPI_ISL_581659\|2020 | ···················· | ···················· | ···················· | ···················· | ···················· | ···················· | |
| L | 6370 hCoV-19/Belgium/ULG-10501/2020\|EPI_ISL_581663\|2020 | ···················· | ···················· | ···················· | ···················· | ···················· | ···················· | |
| L | 6374 hCoV-19/Belgium/ULG-10505/2020\|EPI_ISL_581667\|2020 | ···················· | ···················· | ···················· | ···················· | ···················· | ···················· | |
| L | 6375 hCoV-19/Switzerland/BE-42216413/2020\|EPI_ISL_58167 | ···················· | ···················· | ···················· | ···················· | ·NN················· | ···················· | |
| L | 6378 hCoV-19/Switzerland/BS-42216821/2020\|EPI_ISL_58167 | ···················· | ···················· | ···················· | ···················· | NNN················· | ···················· | 1 |
| L | 6379 hCoV-19/Switzerland/BL-42219090/2020\|EPI_ISL_58167 | ···················· | ···················· | ···················· | ···················· | ···················· | ···················· | |
| L | 6380 hCoV-19/Switzerland/BS-42220323/2020\|EPI_ISL_58167 | ···················· | ···················· | ···················· | ···················· | ···················· | ···················· | |
| L | 6381 hCoV-19/Switzerland/BS-42222049/2020\|EPI_ISL_58168 | ···················· | ···················· | ···················· | ···················· | ···················· | ···················· | |
| L | 6382 hCoV-19/Switzerland/BS-42222240/2020\|EPI_ISL_58168 | ···················· | ···················· | ···················· | ···················· | ···················· | ···················· | |
| L | 6383 hCoV-19/Switzerland/BS-42222420/2020\|EPI_ISL_58168 | ···················· | ···················· | ···················· | ···················· | ···················· | ···················· | |
| L | 6384 hCoV-19/Switzerland/SO-42222503/2020\|EPI_ISL_58168 | ···················· | ···················· | ···················· | ···················· | ···················· | ···················· | |
| L | 6385 hCoV-19/Switzerland/BL-42222531/2020\|EPI_ISL_58168 | ···················· | ···················· | ···················· | ···················· | ···················· | ···················· | |
| L | 6386 hCoV-19/Switzerland/BS-42224191/2020\|EPI_ISL_58168 | ···················· | ·A·················· | ···················· | ···················· | ···················· | ···················· | |
| L | 6387 hCoV-19/Switzerland/BL-42225118/2020\|EPI_ISL_58168 | ···················· | ···················· | ···················· | ···················· | ···················· | ···················· | |
| L | 6388 hCoV-19/Switzerland/BS-42225323/2020\|EPI_ISL_58169 | ···················· | ···················· | ···················· | ···················· | ···················· | ···················· | |
| L | 6389 hCoV-19/USA/CA-CZB-6719/2020\|EPI_ISL_582980\|2020-0 | ···················· | ···················· | ···················· | ···················· | ···················· | ···················· | |
| L | 6390 hCoV-19/USA/CA-CZB-6720/2020\|EPI_ISL_582981\|2020-0 | ···················· | ···················· | ···················· | ···················· | ···················· | ···················· | |
| L | 6391 hCoV-19/USA/CA-CZB-6721/2020\|EPI_ISL_582982\|2020-0 | ···················· | ···················· | ···················· | ···················· | ···················· | ···················· | |
| L | 6392 hCoV-19/USA/CA-CZB-6722/2020\|EPI_ISL_582983\|2020-0 | ···················· | ···················· | ···················· | ···················· | ···················· | ···················· | |
| L | 6393 hCoV-19/USA/CA-CZB-6725/2020\|EPI_ISL_582984\|2020-0 | ···················· | ·A·················· | ···················· | ···················· | ···················· | ···················· | |
| L | 6394 hCoV-19/USA/CA-CZB-6726/2020\|EPI_ISL_582985\|2020-0 | ···················· | ·A·················· | ···················· | ···················· | ···················· | ···················· | |
| L | 6395 hCoV-19/USA/CA-CZB-6727/2020\|EPI_ISL_582986\|2020-0 | ···················· | ·A·················· | ···················· | ···················· | ···················· | ···················· | |
| L | 6396 hCoV-19/USA/CA-CZB-6728/2020\|EPI_ISL_582987\|2020-0 | ···················· | ···················· | ···················· | ···················· | ···················· | ···················· | |
| L | 6397 hCoV-19/USA/CA-CZB-6729/2020\|EPI_ISL_582988\|2020-0 | ···················· | ·A·················· | ···················· | ···················· | ···················· | ···················· | |
| L | 6398 hCoV-19/USA/CA-CZB-6731/2020\|EPI_ISL_582989\|2020-0 | ···················· | ·A·················· | ···················· | ···················· | ···················· | ···················· | |
| L | 6399 hCoV-19/USA/CA-CZB-6732/2020\|EPI_ISL_582990\|2020-0 | ···················· | ···················· | ···················· | ···················· | ···················· | ···················· | |
| L | 6400 hCoV-19/USA/CA-CZB-6745/2020\|EPI_ISL_582999\|2020-0 | ···················· | ···················· | ···················· | ···················· | ···················· | ···················· | |
| L | 6401 hCoV-19/England/MILK-973227/2020\|EPI_ISL_575800\|20 | ···················· | ···················· | ···················· | ···················· | ···················· | ···················· | |
| L | 6402 hCoV-19/Scotland/MILK-974736/2020\|EPI_ISL_575802\|2 | ···················· | ·A·················· | ···················· | ···················· | ···················· | ···A················ | |
| L | 6403 hCoV-19/England/MILK-973096/2020\|EPI_ISL_575803\|20 | ···················· | ···················· | ···················· | ···················· | ···················· | ···················· | |
| L | 6404 hCoV-19/England/MILK-973430/2020\|EPI_ISL_575804\|20 | ···················· | ···················· | ···················· | ···················· | ···················· | ···················· | |
| L | 6405 hCoV-19/England/MILK-97645B/2020\|EPI_ISL_575806\|20 | ···················· | ···················· | ···················· | ···················· | ···················· | ···················· | |
| L | 6406 hCoV-19/England/MILK-976558/2020\|EPI_ISL_575807\|20 | ···················· | ···················· | ···················· | ···················· | ···················· | ···················· | |
| L | 6407 hCoV-19/England/MILK-976673/2020\|EPI_ISL_575809\|20 | ···················· | ···················· | ···················· | ···················· | ···················· | ···················· | |
| L | 6408 hCoV-19/Netherlands/ZH-EMC-572/2020\|EPI_ISL_578000 | ···················· | ···················· | ···················· | ···················· | ···················· | ···················· | |
| L | 6409 hCoV-19/Netherlands/ZH-EMC-573/2020\|EPI_ISL_578001 | ···················· | ···················· | ···················· | ···················· | ···················· | ···················· | |
| L | 6410 hCoV-19/Netherlands/ZH-EMC-574/2020\|EPI_ISL_578002 | ···················· | ···················· | ···················· | ···················· | ······T············· | ···················· | |
| L | 6411 hCoV-19/Netherlands/ZH-EMC-576/2020\|EPI_ISL_578004 | ···················· | G··················· | ···················· | ···················· | ···················· | ···················· | |

```
L 6412 hCoV-19/Netherlands/ZH-EMC-577/2020|EPI_ISL_578005 ···················· ···················· ···················· ···················· ···················· ····················
L 6413 hCoV-19/Netherlands/ZH-EMC-578/2020|EPI_ISL_578006 ···················· ···················· ···················· ···················· ···················· ····················
L 6414 hCoV-19/Netherlands/ZH-EMC-579/2020|EPI_ISL_578007 ···················· ···················· ···················· ···················· ···················· ····················
L 6415 hCoV-19/Netherlands/ZH-EMC-602/2020|EPI_ISL_578030 ···················· ···················· ···················· ···················· ···················· ····················
L 6416 hCoV-19/Netherlands/ZH-EMC-607/2020|EPI_ISL_578035 ···················· ···················· ···················· ···················· ···················· ····················
L 6417 hCoV-19/Netherlands/ZH-EMC-608/2020|EPI_ISL_578036 ···················· ···················· ···················· ···················· ···················· ····················
L 6418 hCoV-19/Netherlands/ZH-EMC-622/2020|EPI_ISL_578050 ···················· ···················· ···················· ···················· ···················· ····················
L 6419 hCoV-19/Netherlands/ZH-EMC-623/2020|EPI_ISL_578051 ···················· ···················· ···················· ···················· ···················· ····················
L 6420 hCoV-19/Netherlands/ZH-EMC-625/2020|EPI_ISL_578053 ···················· ···················· ···················· ···················· ···················· ····················
L 6421 hCoV-19/Netherlands/ZH-EMC-626/2020|EPI_ISL_578054 ···················· ···················· ···················· ···················· ···················· ····················
L 6422 hCoV-19/Netherlands/ZH-EMC-629/2020|EPI_ISL_578057 ···················· ···················· ···················· ···················· ···················· ····················
L 6423 hCoV-19/Netherlands/ZH-EMC-631/2020|EPI_ISL_578059 ···················· ···················· ···················· ···················· ······T············· ····················
L 6424 hCoV-19/Netherlands/ZH-EMC-632/2020|EPI_ISL_578060 ···················· ···················· ···················· ···················· ···················· ····················
L 6425 hCoV-19/Netherlands/ZH-EMC-633/2020|EPI_ISL_578061 ···················· ···················· ···················· ···················· ···················· ····················
L 6426 hCoV-19/Netherlands/ZH-EMC-634/2020|EPI_ISL_578062 ···················· ···················· ···················· ···················· ···················· ····················
L 6427 hCoV-19/Netherlands/ZH-EMC-635/2020|EPI_ISL_578063 ···················· ···················· ···················· ···················· ···················· ····················
L 6428 hCoV-19/Netherlands/ZH-EMC-636/2020|EPI_ISL_578064 ···················· ···················· ···················· ···················· ···················· ····················
L 6429 hCoV-19/Netherlands/ZH-EMC-637/2020|EPI_ISL_578065 ···················· ···················· ···················· ···················· ···················· ····················
L 6430 hCoV-19/Netherlands/ZH-EMC-639/2020|EPI_ISL_578067 ···················· ···················· ···················· ···················· ···················· ····················
L 6431 hCoV-19/Netherlands/ZH-EMC-640/2020|EPI_ISL_578068 ···················· ···················· ···················· ···················· ···················· ····················
L 6432 hCoV-19/Netherlands/ZH-EMC-641/2020|EPI_ISL_578069 ···················· ···················· ···················· ···················· ···················· ····················
L 6433 hCoV-19/Netherlands/ZH-EMC-642/2020|EPI_ISL_578070 ···················· ······A············· ···················· ···················· ···················· ····················
L 6434 hCoV-19/Netherlands/ZH-EMC-643/2020|EPI_ISL_578071 ···················· ···················· ···················· ···················· ···················· ····················
L 6435 hCoV-19/Netherlands/ZH-EMC-644/2020|EPI_ISL_578072 ···················· ····A··············· ···················· ···················· ···················· ····················
L 6436 hCoV-19/Netherlands/ZH-EMC-645/2020|EPI_ISL_578073 ···················· G··················· ···················· ···················· ···················· ····················
L 6437 hCoV-19/Netherlands/ZH-EMC-646/2020|EPI_ISL_578074 ···················· ···················· ···················· ···················· ···················· ····················
L 6438 hCoV-19/Netherlands/ZH-EMC-649/2020|EPI_ISL_578077 ···················· ···················· ···················· ···················· ···················· ····················
L 6439 hCoV-19/Netherlands/ZH-EMC-650/2020|EPI_ISL_578078 ···················· ···················· ···················· ···················· ···················· ····················
L 6440 hCoV-19/Netherlands/ZH-EMC-651/2020|EPI_ISL_578079 ···················· ···················· ···················· ···················· ···················· ····················
L 6441 hCoV-19/India/WB-IICB-006/2020|EPI_ISL_578080|2020 ···················· ···················· ···················· ···················· ···················· ····················
L 6442 hCoV-19/India/WB-IICB-007/2020|EPI_ISL_578081|2020 ···················· ·A·················· ···················· ···················· ···················· ····················
L 6443 hCoV-19/USA/MI-UM-10033834090/2020|EPI_ISL_578089| ···················· ················T··· ···················· ···················· ···················· ····················
L 6444 hCoV-19/New Zealand/20VR2561/2020|EPI_ISL_579300|2 ···················· ···················· ···················· ···················· ···················· ····················
L 6445 hCoV-19/New Zealand/20VR2570/2020|EPI_ISL_579301|2 ···················· ···················· ···················· ···················· ···················· ····················
L 6446 hCoV-19/New Zealand/20VR2571/2020|EPI_ISL_579302|2 ···················· ···················· ···················· ···················· ···················· ····················
L 6448 hCoV-19/New Zealand/20VR2573/2020|EPI_ISL_579304|2 ···················· ···················· ···················· ···················· ···················· ····················
L 6449 hCoV-19/New Zealand/20VR2574/2020|EPI_ISL_579305|2 ···················· ···················· ···················· ···················· ···················· ····················
L 6451 hCoV-19/New Zealand/20VR2576/2020|EPI_ISL_579307|2 ···················· ···················· ···················· ···················· ···················· ····················
L 6454 hCoV-19/New Zealand/20VR2579/2020|EPI_ISL_579310|2 ···················· ···················· ···················· ···················· ···················· ····················
L 6455 hCoV-19/New Zealand/20VR2585/2020|EPI_ISL_579313|2 ···················· ···················· ···················· ···················· ···················· ····················
L 6456 hCoV-19/New Zealand/20VR2586/2020|EPI_ISL_579314|2 ···················· ···················· ···················· ···················· ···················· ····················
L 6458 hCoV-19/New Zealand/20VR2669/2020|EPI_ISL_579330|2 ···················· ···················· ···················· ···················· ···················· ····················
L 6459 hCoV-19/New Zealand/20VR2711/2020|EPI_ISL_579332|2 ···················· ···················· ···················· ···················· ···················· ····················
L 6467 hCoV-19/New Zealand/20VR2942/2020|EPI_ISL_579363|2 ···················· ···················· ···················· ···················· ···················· ····················
L 6470 hCoV-19/New Zealand/20VR3008/2020|EPI_ISL_579380|2 ···················· ·············A······ ···················· ···················· ···················· ····················
L 6472 hCoV-19/New Zealand/20VR3010/2020|EPI_ISL_579382|2 ···················· ···················· ···················· ···················· ···················· ····················
L 6473 hCoV-19/New Zealand/20VR3011/2020|EPI_ISL_579383|2 ···················· ···················· ···················· ···················· ···················· ····················
L 6475 hCoV-19/New Zealand/20VR3014/2020|EPI_ISL_579385|2 ···················· ···················· ···················· ···················· ···················· ····················
L 6477 hCoV-19/New Zealand/20VR3019/2020|EPI_ISL_579388|2 ···················· ·············A······ ···················· ···················· ···················· ····················
L 6478 hCoV-19/New Zealand/20VR3022/2020|EPI_ISL_579389|2 ···················· ···················· ···················· ···················· ···················· ····················
L 6479 hCoV-19/New Zealand/20VR3024/2020|EPI_ISL_579390|2 ···················· ···················· ···················· ···················· ···················· ····················
L 6481 hCoV-19/New Zealand/20VR3026/2020|EPI_ISL_579392|2 ···················· ···················· ···················· ···················· ···················· ····················
L 6486 hCoV-19/New Zealand/20VR3036/2020|EPI_ISL_579398|2 ···················· ···················· ···················· ···················· ···················· ····················
L 6487 hCoV-19/New Zealand/20VR3039/2020|EPI_ISL_579399|2 ···················· ···················· ···················· ···················· ···················· ····················
L 6488 hCoV-19/England/CAMC-9EC511/2020|EPI_ISL_580301|20 ···················· ···················· ···················· ···················· ···················· ····················
L 6489 hCoV-19/England/ALDP-9E66E7/2020|EPI_ISL_580302|20 ···················· ···················· ···················· ···················· ···················· ····················
L 6490 hCoV-19/England/ALDP-9E6D1F/2020|EPI_ISL_580303|20 ···················· ···················· ···················· ···················· ···················· ····················
L 6491 hCoV-19/England/ALDP-9E6CE5/2020|EPI_ISL_580304|20 ···················· ···················· ···················· ···················· ···················· ····················
L 6492 hCoV-19/England/CAMC-9EC599/2020|EPI_ISL_580305|20 ···················· ···················· ···················· ···················· ···················· ····················
L 6493 hCoV-19/England/ALDP-9E677B/2020|EPI_ISL_580306|20 ···················· ·A·················· ···················· ···················· ···················· ···A················
L 6494 hCoV-19/England/ALDP-9E6D00/2020|EPI_ISL_580307|20 ···················· ···················· ···················· ···················· ···················· ····················
L 6495 hCoV-19/England/ALDP-9E6F46/2020|EPI_ISL_580308|20 ···················· ···················· ···················· ···················· ···················· ····················
L 6496 hCoV-19/England/ALDP-9E6C21/2020|EPI_ISL_580309|20 ···················· ···················· ···················· ···················· ···················· ····················
L 6497 hCoV-19/England/ALDP-9E6DD3/2020|EPI_ISL_580310|20 ···················· ···················· ···················· ···················· ···················· ····················
L 6498 hCoV-19/England/ALDP-9E7446/2020|EPI_ISL_580311|20 ···················· ···················· ···················· ···················· ···················· ····················
L 6499 hCoV-19/England/CAMC-9EC6A5/2020|EPI_ISL_580313|20 ···················· ···················· ···················· ···················· ···················· ····················
L 6500 hCoV-19/England/ALDP-9E67B7/2020|EPI_ISL_580314|20 ···················· ·A·················· ···················· ···················· ···················· ····················
L 6501 hCoV-19/England/ALDP-9E627A/2020|EPI_ISL_580315|20 ···················· ···················· ···················· ···················· ···················· ····················
L 6502 hCoV-19/England/ALDP-9E70F7/2020|EPI_ISL_580316|20 ···················· ···················· ···················· ···················· ···················· ····················
L 6503 hCoV-19/England/ALDP-9E72D3/2020|EPI_ISL_580317|20 ···················· ···················· ···················· ···················· ···················· ····················
L 6504 hCoV-19/England/ALDP-9E667E/2020|EPI_ISL_580318|20 ···················· ···················· ···················· ···················· ···················· ····················
L 6505 hCoV-19/England/ALDP-9E68F0/2020|EPI_ISL_580319|20 ···················· ···················· ···················· ···················· ····T··············· ····················
L 6506 hCoV-19/England/ALDP-9E6508/2020|EPI_ISL_580337|20 ···················· ···················· ···················· ···················· ···················· ····················
L 6507 hCoV-19/England/CAMC-9EC06B/2020|EPI_ISL_580338|20 ···················· ···················· ···················· ···················· ···················· ····················
L 6508 hCoV-19/England/ALDP-9E6B06/2020|EPI_ISL_580339|20 ···················· ···················· ···················· ···················· ···················· ····················
L 6509 hCoV-19/England/CAMC-9EC380/2020|EPI_ISL_580362|20 ···················· ···················· ···················· ···················· ···················· ····················
L 6510 hCoV-19/England/ALDP-9E6DB5/2020|EPI_ISL_580363|20 ···················· ···················· ···················· ···················· ···················· ····················
L 6511 hCoV-19/England/CAMC-9EC793/2020|EPI_ISL_580364|20 ···················· ···················· ···················· ···················· ···················· ····················
L 6512 hCoV-19/England/ALDP-9E6E0D/2020|EPI_ISL_580365|20 ···················· ···················· ···················· ···················· ···················· ····················
L 6513 hCoV-19/England/ALDP-9E71A9/2020|EPI_ISL_580366|20 ···················· ···················· ···················· ···················· ···················· ····················
```

| | | | | | | | |
|---|---|---|---|---|---|---|---|
| L | 6514 hCoV-19/England/CAMC-9EC0B6/2020\|EPI_ISL_580367\|20 | ···················· | ···················· | ···················· | ···················· | ···················· | ···················· |
| L | 6515 hCoV-19/England/CAMC-9EC177/2020\|EPI_ISL_580368\|20 | ···················· | ···················· | ···················· | ···················· | ···················· | ···················· |
| L | 6516 hCoV-19/England/CAMC-9EC5E4/2020\|EPI_ISL_580369\|20 | ···················· | ·A·················· | ···················· | ···················· | ···················· | ··A················· |
| L | 6517 hCoV-19/England/ALDP-9E6C03/2020\|EPI_ISL_580370\|20 | ···················· | ···················· | ···················· | ···················· | ···················· | ···················· |
| L | 6518 hCoV-19/England/CAMC-9EC3EA/2020\|EPI_ISL_580371\|20 | ···················· | ···················· | ···················· | ···················· | ···················· | ···················· |
| L | 6519 hCoV-19/England/ALDP-9E7112/2020\|EPI_ISL_580372\|20 | ···················· | ···················· | ···················· | ···················· | ···················· | ···················· |
| L | 6520 hCoV-19/England/CAMC-9EC72A/2020\|EPI_ISL_580373\|20 | ···················· | ···················· | ···················· | ···················· | ···················· | ···················· |
| L | 6521 hCoV-19/England/ALDP-9E6F73/2020\|EPI_ISL_580374\|20 | ···················· | ···················· | ···················· | ···················· | ···················· | ···················· |
| L | 6522 hCoV-19/England/ALDP-9E60BC/2020\|EPI_ISL_580375\|20 | ···················· | ···················· | ···················· | ···················· | ···················· | ···················· |
| L | 6523 hCoV-19/England/ALDP-9E6BCA/2020\|EPI_ISL_580376\|20 | ···················· | ···················· | ···················· | ···················· | ···················· | ···················· |
| L | 6524 hCoV-19/England/ALDP-9E6377/2020\|EPI_ISL_580377\|20 | ···················· | ···················· | ···················· | ···················· | ···················· | ···················· |
| L | 6525 hCoV-19/England/ALDP-9E626B/2020\|EPI_ISL_580378\|20 | ···················· | ···················· | ···················· | ···················· | ···················· | ···················· |
| L | 6526 hCoV-19/England/ALDP-9E632C/2020\|EPI_ISL_580379\|20 | ···················· | ···················· | ···················· | ···················· | ···················· | ···················· |
| L | 6527 hCoV-19/England/ALDP-9E6E2B/2020\|EPI_ISL_580380\|20 | ···················· | ···················· | ···················· | ···················· | ···················· | ···················· |
| L | 6528 hCoV-19/England/ALDP-9E60E9/2020\|EPI_ISL_580381\|20 | ···················· | ···················· | ···················· | ···················· | ···················· | ···················· |
| L | 6529 hCoV-19/England/ALDP-9E7033/2020\|EPI_ISL_580382\|20 | ···················· | ···················· | ···················· | ···················· | ···················· | ···················· |
| L | 6530 hCoV-19/England/ALDP-9E633B/2020\|EPI_ISL_580383\|20 | ···················· | ···················· | ···················· | ···················· | ···················· | ···················· |
| L | 6531 hCoV-19/England/ALDP-9E6896/2020\|EPI_ISL_580384\|20 | ···················· | ···················· | ···················· | ···················· | ···················· | ···················· |
| L | 6532 hCoV-19/England/CAMC-9EC11D/2020\|EPI_ISL_580385\|20 | ···················· | ···················· | ···················· | ···················· | ···················· | ···················· |
| L | 6533 hCoV-19/England/ALDP-9E74DD/2020\|EPI_ISL_580386\|20 | ···················· | ···················· | ···················· | ···················· | ···················· | ···················· |
| L | 6534 hCoV-19/England/ALDP-9E6F82/2020\|EPI_ISL_580387\|20 | ···················· | ···················· | ···················· | ···················· | ···················· | ···················· |
| L | 6535 hCoV-19/England/ALDP-9E6CF4/2020\|EPI_ISL_580388\|20 | ···················· | ···················· | ···················· | ···················· | ···················· | ···················· |
| L | 6536 hCoV-19/England/ALDP-9E68B4/2020\|EPI_ISL_580389\|20 | ···················· | ···················· | ···················· | ···················· | ···················· | ···················· |
| L | 6537 hCoV-19/England/ALDP-9E70D9/2020\|EPI_ISL_580390\|20 | ···················· | ···················· | ···················· | ···················· | ···················· | ···················· |
| L | 6538 hCoV-19/England/CAMC-9EC6C3/2020\|EPI_ISL_580391\|20 | ···················· | ···················· | ···················· | ···················· | ···················· | ···················· |
| L | 6539 hCoV-19/England/ALDP-9E6ADC/2020\|EPI_ISL_580392\|20 | ···················· | ···················· | ···················· | ···················· | ···················· | ···················· |
| L | 6540 hCoV-19/England/CAMC-9EC335/2020\|EPI_ISL_580393\|20 | ···················· | ···················· | ···················· | ···················· | ···················· | ···················· |
| L | 6541 hCoV-19/England/CAMC-9EC14A/2020\|EPI_ISL_580394\|20 | ···················· | ·A·················· | ···················· | ···················· | ···················· | ··A················· |
| L | 6542 hCoV-19/England/ALDP-9E6887/2020\|EPI_ISL_580395\|20 | ···················· | ···················· | ···················· | ···················· | ···················· | ···················· |
| L | 6543 hCoV-19/England/CAMC-9EC0F2/2020\|EPI_ISL_580396\|20 | ···················· | ···················· | ···················· | ···················· | ···················· | ···················· |
| L | 6544 hCoV-19/England/ALDP-9E61C8/2020\|EPI_ISL_580397\|20 | ···················· | ···················· | ···················· | ···················· | ···················· | ···················· |
| L | 6545 hCoV-19/England/ALDP-9E67C6/2020\|EPI_ISL_580398\|20 | ···················· | ·A·················· | ···················· | ···················· | ···················· | ···················· |
| L | 6546 hCoV-19/England/ALDP-9E72B5/2020\|EPI_ISL_580399\|20 | ···················· | ···················· | ···················· | ···················· | ···················· | ···················· |
| L | 6547 hCoV-19/Belgium/ULG-10418/2020\|EPI_ISL_581600\|2020 | ···················· | ···················· | ···················· | ···················· | ···················· | ···················· |
| L | 6548 hCoV-19/Belgium/ULG-10419/2020\|EPI_ISL_581601\|2020 | ···················· | ···················· | ···················· | ···················· | ···················· | ···················· |
| L | 6549 hCoV-19/Belgium/ULG-10420/2020\|EPI_ISL_581602\|2020 | ···················· | ···················· | ···················· | ···················· | ···················· | ···················· |
| L | 6550 hCoV-19/Belgium/ULG-10421/2020\|EPI_ISL_581603\|2020 | ···················· | ···················· | ···················· | ···················· | ···················· | ···················· |
| L | 6551 hCoV-19/Belgium/ULG-10422/2020\|EPI_ISL_581604\|2020 | ···················· | ···················· | ···················· | ···················· | ···················· | ···················· |
| L | 6552 hCoV-19/Belgium/ULG-10425/2020\|EPI_ISL_581605\|2020 | ···················· | ···················· | ···················· | ···················· | ···················· | ···················· |
| L | 6553 hCoV-19/Belgium/ULG-10426/2020\|EPI_ISL_581606\|2020 | ···················· | ···················· | ···················· | ···················· | ···················· | ···················· |
| L | 6554 hCoV-19/Belgium/ULG-10429/2020\|EPI_ISL_581607\|2020 | ···················· | ···················· | ···················· | ···················· | ···················· | ···················· |
| L | 6555 hCoV-19/Belgium/ULG-10430/2020\|EPI_ISL_581608\|2020 | ···················· | ···················· | ···················· | ···················· | ···················· | ···················· |
| L | 6556 hCoV-19/Belgium/ULG-10431/2020\|EPI_ISL_581609\|2020 | ···················· | ···················· | ···················· | ···················· | ···················· | ···················· |
| L | 6557 hCoV-19/Belgium/ULG-10432/2020\|EPI_ISL_581610\|2020 | ···················· | ···················· | ···················· | ···················· | ···················· | ···················· |
| L | 6558 hCoV-19/Belgium/ULG-10433/2020\|EPI_ISL_581611\|2020 | ···················· | ···················· | ···················· | ···················· | ···················· | ···················· |
| L | 6559 hCoV-19/Belgium/ULG-10434/2020\|EPI_ISL_581612\|2020 | ···················· | ···················· | ···················· | ···················· | ···················· | ···················· |
| L | 6560 hCoV-19/Belgium/ULG-10440/2020\|EPI_ISL_581617\|2020 | ···················· | ···················· | ···················· | ···················· | ···················· | ···················· |
| L | 6561 hCoV-19/Belgium/ULG-10441/2020\|EPI_ISL_581618\|2020 | ···················· | ···················· | ···················· | ···················· | ···················· | ···················· |
| L | 6562 hCoV-19/Belgium/ULG-10442/2020\|EPI_ISL_581619\|2020 | ···················· | ···················· | ···················· | ···················· | ···················· | ···················· |
| L | 6563 hCoV-19/Belgium/ULG-10457/2020\|EPI_ISL_581630\|2020 | ···················· | ···················· | ···················· | ···················· | ···················· | ···················· |
| L | 6564 hCoV-19/Belgium/ULG-10458/2020\|EPI_ISL_581631\|2020 | ···················· | ···················· | ···················· | ···················· | ···················· | ···················· |
| L | 6565 hCoV-19/Belgium/ULG-10461/2020\|EPI_ISL_581632\|2020 | ···················· | ···················· | ···················· | ···················· | ···················· | ···················· |
| L | 6566 hCoV-19/Belgium/ULG-10462/2020\|EPI_ISL_581633\|2020 | ···················· | ···················· | ···················· | ···················· | ···················· | ···················· |
| L | 6567 hCoV-19/Belgium/ULG-10463/2020\|EPI_ISL_581634\|2020 | ···················· | ···················· | ···················· | ···················· | ···················· | ···················· |
| L | 6568 hCoV-19/Belgium/ULG-10464/2020\|EPI_ISL_581635\|2020 | ···················· | ···················· | ···················· | ···················· | ···················· | ···················· |
| L | 6572 hCoV-19/Belgium/ULG-10469/2020\|EPI_ISL_581639\|2020 | ···················· | ···················· | ···················· | ···················· | ···················· | ···················· |
| L | 6577 hCoV-19/Belgium/ULG-10475/2020\|EPI_ISL_581644\|2020 | ···················· | ···················· | ···················· | ···················· | ···················· | ···················· |
| L | 6581 hCoV-19/Belgium/ULG-10483/2020\|EPI_ISL_581648\|2020 | ···················· | ···················· | ···················· | ···················· | ···················· | ···················· |
| L | 6583 hCoV-19/Belgium/ULG-10498/2020\|EPI_ISL_581660\|2020 | ···················· | ···················· | ···················· | ···················· | ···················· | ···················· |
| L | 6584 hCoV-19/Switzerland/BS-42216174/2020\|EPI_ISL_58166 | ···················· | ···················· | ···················· | ···················· | ···················· | ···················· |
| L | 6585 hCoV-19/Switzerland/BS-42216240/2020\|EPI_ISL_58166 | ···················· | ···················· | ···················· | ···················· | ·NN················· | ···················· |
| L | 6586 hCoV-19/Switzerland/BS-42227559/2020\|EPI_ISL_58169 | ···················· | ···················· | ···················· | ···················· | ···················· | ···················· |
| L | 6587 hCoV-19/Switzerland/BS-42227598/2020\|EPI_ISL_58169 | ···················· | ···················· | ···················· | ···················· | ···················· | ···················· |
| L | 6588 hCoV-19/Switzerland/BS-42227651/2020\|EPI_ISL_58169 | ···················· | ·A·················· | ···················· | ···················· | ···················· | ···················· |
| L | 6589 hCoV-19/Switzerland/BS-42227717/2020\|EPI_ISL_58169 | ···················· | ···················· | ···················· | ···················· | ···················· | ···················· |
| L | 6590 hCoV-19/Switzerland/BL-42227720/2020\|EPI_ISL_58169 | ···················· | ···················· | ···················· | ···················· | ···················· | ···················· |
| L | 6591 hCoV-19/Switzerland/JU-42228682/2020\|EPI_ISL_58169 | ···················· | ···················· | ···················· | ···················· | ···················· | ···················· |
| L | 6592 hCoV-19/USA/CA-CZB-6965/2020\|EPI_ISL_582900\|2020-0 | ···················· | ···················· | ···················· | ···················· | ···················· | ···················· |
| L | 6593 hCoV-19/USA/CA-CZB-6966/2020\|EPI_ISL_582901\|2020-0 | ···················· | ·A·················· | ···················· | ···················· | ···················· | ···················· |
| L | 6594 hCoV-19/USA/CA-CZB-6967/2020\|EPI_ISL_582902\|2020-0 | ···················· | ·A·················· | ···················· | ···················· | ···················· | ···················· |
| L | 6595 hCoV-19/USA/CA-CZB-6968/2020\|EPI_ISL_582903\|2020-0 | ···················· | ·A·················· | ···················· | ···················· | ···················· | ···················· |
| L | 6596 hCoV-19/USA/CA-CZB-6971/2020\|EPI_ISL_582904\|2020-0 | ···················· | ·A·················· | ···················· | ···················· | ···················· | ···················· |
| L | 6597 hCoV-19/USA/CA-CZB-6973/2020\|EPI_ISL_582905\|2020-0 | ···················· | ·A·················· | ···················· | ···················· | ···················· | ···················· |
| L | 6598 hCoV-19/USA/CA-CZB-6974/2020\|EPI_ISL_582906\|2020-0 | ···················· | ·A·················· | ···················· | ···················· | ···················· | ···················· |
| L | 6599 hCoV-19/USA/CA-CZB-6975/2020\|EPI_ISL_582907\|2020-0 | ···················· | ·A·················· | ···················· | ···················· | ···················· | ···················· |
| L | 6600 hCoV-19/USA/CA-CZB-6978/2020\|EPI_ISL_582908\|2020-0 | ···················· | ·A·················· | ···················· | ···················· | ···················· | ··A················· |
| L | 6601 hCoV-19/USA/CA-CZB-6981/2020\|EPI_ISL_582909\|2020-0 | ···················· | ·A·················· | ···················· | ···················· | ···················· | ···················· |
| L | 6602 hCoV-19/USA/CA-CZB-6983/2020\|EPI_ISL_582910\|2020-0 | ···················· | ·A·················· | ···················· | ···················· | ···················· | ···················· |
| L | 6603 hCoV-19/USA/CA-CZB-6984/2020\|EPI_ISL_582911\|2020-0 | ···················· | ·A·················· | ···················· | ···················· | ···················· | ···················· |
| L | 6604 hCoV-19/USA/CA-CZB-6985/2020\|EPI_ISL_582912\|2020-0 | ···················· | ·A·················· | ···················· | ···················· | ···················· | ···················· |

L 6605 hCoV-19/USA/CA-CZB-6986/2020|EPI_ISL_582913|2020-0 ···················· ·A·················· ···················· ···················· ···················· ····················
L 6606 hCoV-19/USA/CA-CZB-6987/2020|EPI_ISL_582914|2020-0 ···················· ···················· ···················· ···················· ···················· ····················
L 6607 hCoV-19/USA/CA-CZB-6988/2020|EPI_ISL_582915|2020-0 ···················· ···················· ···················· ···················· ···················· ····················
L 6608 hCoV-19/USA/CA-CZB-6989/2020|EPI_ISL_582916|2020-0 ···················· ···················· ···················· ···················· ···················· ····················
L 6609 hCoV-19/USA/CA-CZB-6990/2020|EPI_ISL_582917|2020-0 ···················· ···················· ···················· ···················· ···················· ····················
L 6610 hCoV-19/USA/CA-CZB-6991/2020|EPI_ISL_582918|2020-0 ···················· ···················· ···················· ···················· ···················· ····················
L 6611 hCoV-19/USA/CA-CZB-6992/2020|EPI_ISL_582919|2020-0 ···················· ···················· ···················· ···················· ···················· ····················
L 6612 hCoV-19/USA/CA-CZB-6994/2020|EPI_ISL_582921|2020-0 ···················· ···················· ···················· ···················· ···················· ····················
L 6613 hCoV-19/USA/CA-CZB-6995/2020|EPI_ISL_582922|2020-0 ···················· ···················· ···················· ···················· ···················· ····················
L 6614 hCoV-19/USA/CA-CZB-6996/2020|EPI_ISL_582923|2020-0 ···················· ···················· ···················· ···················· ···················· ····················
L 6615 hCoV-19/USA/CA-CZB-6997/2020|EPI_ISL_582924|2020-0 ···················· ···················· ···················· ···················· ···················· ····················
L 6616 hCoV-19/USA/CA-CZB-6999/2020|EPI_ISL_582925|2020-0 ···················· ···················· ···················· ···················· ···················· ····················
L 6617 hCoV-19/USA/CA-CZB-7000/2020|EPI_ISL_582926|2020-0 ···················· ···················· ···················· ···················· ···················· ····················
L 6618 hCoV-19/USA/CA-CZB-7003/2020|EPI_ISL_582928|2020-0 ············T······· ···················· ···················· ···················· ···················· ····················
L 6619 hCoV-19/USA/CA-CZB-7145/2020|EPI_ISL_582929|2020-0 ···················· ·A·················· ···················· ···················· ···················· ····················
L 6620 hCoV-19/USA/CA-CZB-7146/2020|EPI_ISL_582930|2020-0 ···················· ·A·················· ···················· ···················· ···················· ····················
L 6621 hCoV-19/USA/CA-CZB-7147/2020|EPI_ISL_582931|2020-0 ···················· ·A·················· ···················· ···················· ···················· ····················
L 6622 hCoV-19/USA/CA-CZB-7148/2020|EPI_ISL_582932|2020-0 ···················· ·A·················· ···················· ···················· ···················· ····················
L 6623 hCoV-19/USA/CA-CZB-7149/2020|EPI_ISL_582933|2020-0 ···················· ·A·················· ···················· ···················· ···················· ····················
L 6624 hCoV-19/USA/CA-CZB-7152/2020|EPI_ISL_582934|2020-0 ···················· ·A·················· ···················· ···················· ···················· ····················
L 6625 hCoV-19/USA/CA-CZB-7153/2020|EPI_ISL_582935|2020-0 ···················· ···················· ···················· ···················· ···················· ····················
L 6626 hCoV-19/USA/CA-CZB-7156/2020|EPI_ISL_582936|2020-0 ···················· ···················· ···················· ···················· ···················· ····················
L 6627 hCoV-19/USA/CA-CZB-7199/2020|EPI_ISL_582937|2020-0 ···················· ···················· ···················· ···················· ···················· ····················
L 6628 hCoV-19/USA/CA-CZB-7202/2020|EPI_ISL_582938|2020-0 ···················· ·A·················· ···················· ···················· ···················· ····················
L 6629 hCoV-19/USA/CA-CZB-7203/2020|EPI_ISL_582939|2020-0 ···················· ···················· ···················· ···················· ···················· ····················
L 6630 hCoV-19/USA/CA-CZB-11007/2020|EPI_ISL_582949|2020- ···················· ···················· ···················· ···················· ···················· ····················
L 6631 hCoV-19/USA/CA-CZB-5833/2020|EPI_ISL_582960|2020-0 ···················· ·A·················· ···················· ···················· ···················· ····················
L 6632 hCoV-19/USA/CA-CZB-5812/2020|EPI_ISL_582961|2020-0 ···················· ···················· ···················· ···················· ···················· ····················
L 6633 hCoV-19/USA/CA-CZB-5780/2020|EPI_ISL_582962|2020-0 ···················· ···················· ···················· ···················· ···················· ····················
L 6634 hCoV-19/USA/CA-CZB-5769/2020|EPI_ISL_582963|2020-0 ···················· ···················· ···················· ···················· ···················· ····················
L 6636 hCoV-19/USA/CA-CZB-5750/2020|EPI_ISL_582965|2020-0 ···················· ···················· ···················· ···················· ···················· ····················
L 6637 hCoV-19/USA/CA-CZB-5749/2020|EPI_ISL_582966|2020-0 ···················· ···················· ···················· ···················· ···················· ····················
L 6638 hCoV-19/USA/CA-CZB-5746/2020|EPI_ISL_582967|2020-0 ···················· ·A·················· ···················· ···················· ···················· ····················
L 6639 hCoV-19/USA/CA-CZB-5743/2020|EPI_ISL_582968|2020-0 ···················· ···················· ···················· ···················· ···················· ····················
L 6640 hCoV-19/USA/CA-CZB-5704/2020|EPI_ISL_582969|2020-0 ···················· ···················· ···················· ···················· ···················· ····················
L 6641 hCoV-19/USA/CA-CZB-5703/2020|EPI_ISL_582970|2020-0 ···················· ···················· ···················· ···················· ···················· ····················
L 6642 hCoV-19/USA/CA-CZB-5698/2020|EPI_ISL_582971|2020-0 ···················· ···················· ···················· ···················· ···················· ····················
L 6643 hCoV-19/USA/CA-CZB-5682/2020|EPI_ISL_582972|2020-0 ···················· ···················· ···················· ···················· ···················· ····················
L 6645 hCoV-19/USA/CA-CZB-5140/2020|EPI_ISL_582974|2020-0 ···················· ·A·················· ···················· ···················· ···················· ····················
L 6646 hCoV-19/USA/CA-CZB-6715/2020|EPI_ISL_582976|2020-0 ···················· ···················· ···················· ···················· ···················· ····················
L 6647 hCoV-19/USA/CA-CZB-6716/2020|EPI_ISL_582977|2020-0 ···················· ·A·················· ···················· ···················· ···················· ····················
L 6648 hCoV-19/USA/CA-CZB-6717/2020|EPI_ISL_582978|2020-0 ···················· ···················· ···················· ···················· ···················· ····················
L 6649 hCoV-19/USA/CA-CZB-6718/2020|EPI_ISL_582979|2020-0 ···················· ···················· ···················· ···················· ···················· ····················
L 6650 hCoV-19/USA/CA-CZB-6733/2020|EPI_ISL_582991|2020-0 ···················· ···················· ···················· ···················· ···················· ····················
L 6651 hCoV-19/USA/CA-CZB-6734/2020|EPI_ISL_582992|2020-0 ···················· ·A····A············· ···················· ···················· ···················· ····················
L 6652 hCoV-19/USA/CA-CZB-6735/2020|EPI_ISL_582993|2020-0 ···················· ·A····A············· ···················· ···················· ···················· ····················
L 6653 hCoV-19/USA/CA-CZB-6736/2020|EPI_ISL_582994|2020-0 ···················· ·A····A············· ···················· ···················· ···················· ····················
L 6654 hCoV-19/USA/CA-CZB-6739/2020|EPI_ISL_582995|2020-0 ···················· ·A·················· ···················· ···················· ···················· ····················
L 6655 hCoV-19/USA/CA-CZB-6740/2020|EPI_ISL_582996|2020-0 ···················· ···················· ···················· ···················· ···················· ····················
L 6656 hCoV-19/USA/CA-CZB-6742/2020|EPI_ISL_582998|2020-0 ···················· ·A·················· ···················· ···················· ···················· ····················
L 6657 hCoV-19/England/CAMC-9C1749/2020|EPI_ISL_573200|20 ···················· ······C············· ···················· ···················· ···················· ····················
L 6658 hCoV-19/England/CAMC-9C1758/2020|EPI_ISL_573201|20 ···················· ···················· ···················· ···················· ···················· ····················
L 6659 hCoV-19/England/CAMC-9C1767/2020|EPI_ISL_573202|20 ···················· ···················· ···················· ···················· ···················· ····················
L 6660 hCoV-19/England/CAMC-9C1794/2020|EPI_ISL_573203|20 ···················· ···················· ···················· ···················· ···················· ····················
L 6661 hCoV-19/England/CAMC-9C180A/2020|EPI_ISL_573204|20 ···················· ···················· ···················· ···················· ···················· ····················
L 6662 hCoV-19/England/CAMC-9C1819/2020|EPI_ISL_573205|20 ···················· ···················· ···················· ···················· ···················· ····················
L 6663 hCoV-19/England/CAMC-9C1837/2020|EPI_ISL_573206|20 ···················· ···················· ···················· ···················· ···················· ····················
L 6664 hCoV-19/England/CAMC-9C1846/2020|EPI_ISL_573207|20 ···················· ···················· ···················· ···················· ···················· ····················
L 6665 hCoV-19/England/CAMC-9C18A0/2020|EPI_ISL_573208|20 ···················· ···················· ···················· ···················· ····T··············· ····················
L 6666 hCoV-19/England/CAMC-9C1961/2020|EPI_ISL_573209|20 ···················· ·A·················· ···················· ···················· ···················· ···A················
L 6667 hCoV-19/England/CAMC-9C1970/2020|EPI_ISL_573210|20 ···················· ···················· ···················· ···················· ···················· ····················
L 6668 hCoV-19/England/CAMC-9C19AD/2020|EPI_ISL_573211|20 ···················· ···················· ···················· ···················· ···················· ····················
L 6669 hCoV-19/England/CAMC-9C1D56/2020|EPI_ISL_573232|20 ···················· ···················· ···················· ···················· ···················· ····················
L 6670 hCoV-19/England/CAMC-9C1D92/2020|EPI_ISL_573234|20 ···················· ···················· ···················· ···················· ···················· ····················
L 6671 hCoV-19/England/CAMC-9C1DDE/2020|EPI_ISL_573235|20 ···················· ···················· ···················· ···················· ···················· ····················
L 6672 hCoV-19/England/LIVE-1D1D3D/2020|EPI_ISL_573236|20 ···················· ···················· ···················· ···················· ···················· ····················
L 6673 hCoV-19/England/LIVE-1D1D6A/2020|EPI_ISL_573237|20 ···················· ···················· ···················· ···················· ···················· ····················
L 6674 hCoV-19/England/LIVE-1D1D79/2020|EPI_ISL_573238|20 ···················· ···················· ···················· ···················· ···················· ····················
L 6675 hCoV-19/England/LIVE-1D1DC4/2020|EPI_ISL_573239|20 ···················· ···················· ···················· ···················· ···················· ····················
L 6676 hCoV-19/England/LIVE-1D1EEF/2020|EPI_ISL_573240|20 ···················· ···················· ···················· ···················· ···················· ····················
L 6677 hCoV-19/England/LIVE-1D2024/2020|EPI_ISL_573241|20 ···················· ···················· ···················· ···················· ···················· ····················
L 6678 hCoV-19/England/LIVE-1D209D/2020|EPI_ISL_573242|20 ···················· ···················· ···················· ···················· ···················· ····················
L 6679 hCoV-19/England/LIVE-1D20E8/2020|EPI_ISL_573243|20 ···················· ···················· ···················· ···················· ···················· ····················
L 6680 hCoV-19/England/LIVE-1D219A/2020|EPI_ISL_573244|20 ···················· ···················· ···················· ···················· ···················· ····················
L 6681 hCoV-19/England/LIVE-1D230D/2020|EPI_ISL_573245|20 ···················· ···················· ···················· ···················· ···················· ····················
L 6682 hCoV-19/England/LIVE-1D6DA1/2020|EPI_ISL_573246|20 ···················· ···················· ···················· ···················· ···················· ····················
L 6683 hCoV-19/England/MILK-9A7F41/2020|EPI_ISL_573247|20 ···················· ···················· ···················· ···················· ···················· ····················
L 6685 hCoV-19/England/MILK-9A7F6F/2020|EPI_ISL_573249|20 ···················· ···················· ···················· ···················· ···················· ····················
L 6687 hCoV-19/England/MILK-9A7FE7/2020|EPI_ISL_573251|20 ···················· ···················· ·········T·········· ···················· ······T············· ····················
L 6688 hCoV-19/England/MILK-9A7FF6/2020|EPI_ISL_573252|20 ···················· ···················· ···················· ···················· ···················· ····················

```
L 6689 hCoV-19/England/MILK-9C4685/2020|EPI_ISL_573253|20 ···················· ···················· ····················· ···················· ···················· ····················
L 6690 hCoV-19/England/MILK-9C4755/2020|EPI_ISL_573254|20 ···················· ···················· ····················· ···················· ···················· ····················
L 6691 hCoV-19/England/MILK-9C4764/2020|EPI_ISL_573255|20 ···················· ···················· ····················· ···················· ···················· ····················
L 6692 hCoV-19/England/MILK-9C47DD/2020|EPI_ISL_573256|20 ···················· ···················· ····················· ···················· ···················· ····················
L 6693 hCoV-19/England/MILK-9C47FB/2020|EPI_ISL_573257|20 ···················· ···················· ····················· ···················· ···················· ····················
L 6694 hCoV-19/England/MILK-9C489E/2020|EPI_ISL_573258|20 ···················· ···················· ····················· ···················· ···················· ····················
L 6695 hCoV-19/England/MILK-9C4913/2020|EPI_ISL_573259|20 ···················· ···················· ····················· ···················· ·····T·············· ····················
L 6696 hCoV-19/England/MILK-9C4931/2020|EPI_ISL_573260|20 ···················· ···················· ····················· ···················· ···················· ····················
L 6697 hCoV-19/England/MILK-9C49E6/2020|EPI_ISL_573261|20 ···················· ···················· ····················· ···················· ···················· ····················
L 6698 hCoV-19/England/MILK-9C4A10/2020|EPI_ISL_573262|20 ···················· ·A·················· ····················· ···················· ···················· ···A················
L 6699 hCoV-19/England/MILK-9C4A3E/2020|EPI_ISL_573263|20 ···················· ···················· ····················· ···················· ···················· ····················
L 6700 hCoV-19/England/MILK-9C4A5C/2020|EPI_ISL_573264|20 ···················· ···················· ····················· ···················· ···················· ····················
L 6701 hCoV-19/England/MILK-9C4B0E/2020|EPI_ISL_573265|20 ···················· ···················· ····················· ···················· ···················· ····················
L 6702 hCoV-19/England/MILK-9C6E12/2020|EPI_ISL_573266|20 ···················· ···················· ····················· ···················· ···················· ····················
L 6703 hCoV-19/England/MILK-9C6E5E/2020|EPI_ISL_573267|20 ···················· ···················· ····················· ···················· ···················· ····················
L 6704 hCoV-19/England/MILK-9C6F1F/2020|EPI_ISL_573268|20 ···················· ···················· ····················· ···················· ···················· ····················
L 6706 hCoV-19/England/MILK-9C6FB5/2020|EPI_ISL_573270|20 ···················· ···················· ····················· ···················· ···················· ····················
L 6707 hCoV-19/England/MILK-9C6FD3/2020|EPI_ISL_573271|20 ···················· ···················· ····················· ···················· ···················· ····················
L 6708 hCoV-19/England/MILK-9C701B/2020|EPI_ISL_573272|20 ···················· ···················· ····················· ···················· ···················· ····················
L 6709 hCoV-19/England/MILK-9C7039/2020|EPI_ISL_573273|20 ···················· ···················· ····················· ···················· ···················· ····················
L 6710 hCoV-19/England/MILK-9C7057/2020|EPI_ISL_573274|20 ···················· ···················· ····················· ···················· ···················· ····················
L 6713 hCoV-19/England/MILK-9C7093/2020|EPI_ISL_573277|20 ···················· ·A·················· ····················· ···················· ···················· ····················
L 6714 hCoV-19/England/MILK-9C70A2/2020|EPI_ISL_573278|20 ···················· ···················· ····················· ···················· ···················· ····················
L 6715 hCoV-19/England/MILK-9C70B1/2020|EPI_ISL_573279|20 ···················· ···················· ····················· ···················· ·····T·············· ····················
L 6716 hCoV-19/England/MILK-9C70DF/2020|EPI_ISL_573280|20 ···················· ···················· ····················· ···················· ···················· ····················
L 6719 hCoV-19/England/MILK-9C7154/2020|EPI_ISL_573283|20 ···················· ···················· ····················· ···················· ···················· ····················
L 6720 hCoV-19/England/MILK-9C7181/2020|EPI_ISL_573284|20 ···················· ···················· ····················· ···················· ···················· ····················
L 6722 hCoV-19/England/MILK-9C71AF/2020|EPI_ISL_573286|20 ···················· ···················· ····················· ···················· ···················· ····················
L 6723 hCoV-19/England/MILK-9C71DC/2020|EPI_ISL_573287|20 ···················· ···················· ····················· ···················· ···················· ····················
L 6724 hCoV-19/England/MILK-9C71FA/2020|EPI_ISL_573288|20 ···················· ···················· ····················· ···················· ···················· ····················
L 6725 hCoV-19/England/MILK-9C7215/2020|EPI_ISL_573289|20 ···················· ·A·················· ····················· ···················· ···················· ···A················
L 6726 hCoV-19/England/MILK-9C7224/2020|EPI_ISL_573290|20 ···················· ···················· ····················· ···················· ···················· ····················
L 6727 hCoV-19/England/MILK-9C7251/2020|EPI_ISL_573291|20 ···················· ···················· ····················· ···················· ···················· ····················
L 6728 hCoV-19/England/MILK-9C727F/2020|EPI_ISL_573292|20 ···················· ···················· ····················· ···················· ···················· ····················
L 6729 hCoV-19/England/MILK-9C72AC/2020|EPI_ISL_573293|20 ···················· ···················· ····················· ···················· ···················· ····················
L 6731 hCoV-19/England/MILK-9C72F7/2020|EPI_ISL_573295|20 ···················· ···················· ····················· ···················· ···················· ····················
L 6732 hCoV-19/England/MILK-9C7303/2020|EPI_ISL_573296|20 ···················· ···················· ····················· ···················· ···················· ····················
L 6733 hCoV-19/England/MILK-9C7312/2020|EPI_ISL_573297|20 ···················· ···················· ····················· ···················· ···················· ····················
L 6734 hCoV-19/England/MILK-9C734F/2020|EPI_ISL_573298|20 ···················· ···················· ····················· ···················· ···················· ····················
L 6735 hCoV-19/England/MILK-9C735E/2020|EPI_ISL_573299|20 ···················· ···················· ····················· ···················· ···················· ····················
L 6736 hCoV-19/Singapore/1015/2020|EPI_ISL_574500|2020-03 ···················· ···················· ····················· ···················· ···················· ····················
L 6737 hCoV-19/Singapore/1016/2020|EPI_ISL_574501|2020-03 ···················· ···················· ····················· ···················· ···················· ····················
L 6738 hCoV-19/Singapore/1017/2020|EPI_ISL_574502|2020-03 ···········T········ ···················· ····················· ···················· ···················· ····················
L 6739 hCoV-19/Singapore/1018/2020|EPI_ISL_574503|2020-09 ···················· ···················· ····················· ···················· ···················· ···A················
L 6740 hCoV-19/Singapore/1019/2020|EPI_ISL_574504|2020-03 ···················· ···················· ····················· ···················· ···················· ····················
L 6741 hCoV-19/Singapore/1020/2020|EPI_ISL_574505|2020-03 ···················· ···················· ····················· ···················· ···················· ····················
L 6742 hCoV-19/Singapore/1021/2020|EPI_ISL_574506|2020-03 ···················· ···················· ····················· ···················· ···················· ····················
L 6743 hCoV-19/Singapore/1022/2020|EPI_ISL_574507|2020-03 ···················· ···················· ····················· ···················· ···················· ····················
L 6744 hCoV-19/Singapore/1023/2020|EPI_ISL_574508|2020-03 ···················· ···················· ····················· ···················· ···················· ····················
L 6745 hCoV-19/Singapore/1024/2020|EPI_ISL_574509|2020-03 ···················· ···················· ····················· ···················· ···················· ····················
L 6746 hCoV-19/Singapore/1025/2020|EPI_ISL_574510|2020-03 ···················· ···················· ····················· ···················· ···················· ····················
L 6747 hCoV-19/Singapore/1026/2020|EPI_ISL_574511|2020-03 ···················· ···················· ····················· ···················· ···················· ····················
L 6748 hCoV-19/Singapore/1027/2020|EPI_ISL_574512|2020-03 ···················· ···················· ····················· ···················· ···················· ····················
L 6749 hCoV-19/Singapore/1028/2020|EPI_ISL_574513|2020-03 ···················· ···················· ····················· ···················· ···················· ····················
L 6750 hCoV-19/Singapore/1029/2020|EPI_ISL_574514|2020-03 ···················· ···················· ····················· ···················· ···················· ····················
L 6751 hCoV-19/Singapore/1030/2020|EPI_ISL_574515|2020-03 ···················· ···················· ····················· ···················· ···················· ····················
L 6752 hCoV-19/Singapore/1031/2020|EPI_ISL_574516|2020-03 ···················· ···················· ····················· ···················· ···················· ····················
L 6753 hCoV-19/Singapore/1032/2020|EPI_ISL_574517|2020-03 ···················· ···················· ····················· ···················· ···················· ····················
L 6754 hCoV-19/Singapore/1033/2020|EPI_ISL_574518|2020-03 ···················· ···················· ····················· ···················· ···················· ····················
L 6755 hCoV-19/Singapore/1034/2020|EPI_ISL_574519|2020-09 ···················· ···················· ····················· ···················· ···················· ····················
L 6756 hCoV-19/Singapore/1035/2020|EPI_ISL_574520|2020-03 ···················· ···················· ····················· ···················· ···················· ····················
L 6757 hCoV-19/Singapore/1036/2020|EPI_ISL_574521|2020-03 ···················· ···················· ····················· ···················· ···················· ····················
L 6758 hCoV-19/Singapore/1037/2020|EPI_ISL_574522|2020-03 ···················· ···················· ····················· ···················· ···················· ····················
L 6759 hCoV-19/Singapore/1038/2020|EPI_ISL_574523|2020-03 ···················· ···················· ····················· ···················· ···················· ····················
L 6760 hCoV-19/Singapore/1039/2020|EPI_ISL_574524|2020-03 ···················· ···················· ····················· ···················· ···················· ····················
L 6761 hCoV-19/Singapore/1040/2020|EPI_ISL_574525|2020-03 ···················· ···················· ····················· ···················· ···················· ····················
L 6762 hCoV-19/Singapore/1041/2020|EPI_ISL_574526|2020-03 ···················· ·············A······ ····················· ···················· ···················· ····················
L 6763 hCoV-19/Singapore/1042/2020|EPI_ISL_574527|2020-03 ···················· ···················· ····················· ···················· ···················· ····················
L 6764 hCoV-19/Singapore/1043/2020|EPI_ISL_574528|2020-03 ···················· ···················· ····················· ···················· ···················· ····················
L 6765 hCoV-19/Singapore/1044/2020|EPI_ISL_574529|2020-03 ···················· ···················· ····················· ···················· ···················· ····················
L 6766 hCoV-19/Singapore/1045/2020|EPI_ISL_574530|2020-03 ···················· ···················· ····················· ···················· ···················· ····················
L 6767 hCoV-19/Singapore/1046/2020|EPI_ISL_574531|2020-03 ···················· ···················· ····················· ···················· ···················· ····················
L 6768 hCoV-19/Singapore/1047/2020|EPI_ISL_574532|2020-03 ···················· ···················· ····················· ···················· ···················· ····················
L 6769 hCoV-19/Singapore/1048/2020|EPI_ISL_574533|2020-03 ···················· ···················· ····················· ···················· ···················· ····················
L 6770 hCoV-19/Singapore/1049/2020|EPI_ISL_574534|2020-03 ···················· ···················· ····················· ···················· ···················· ····················
L 6771 hCoV-19/Singapore/1050/2020|EPI_ISL_574535|2020-03 ···················· ···················· ····················· ···················· ···················· ····················
L 6772 hCoV-19/Singapore/1051/2020|EPI_ISL_574536|2020-03 ···················· ···················· ····················· ···················· ···················· ····················
L 6773 hCoV-19/Singapore/1052/2020|EPI_ISL_574537|2020-03 ···················· ···················· ····················· ···················· ···················· ····················
L 6774 hCoV-19/Singapore/1053/2020|EPI_ISL_574538|2020-03 ···················· ···················· ····················· ···················· ···················· ····················
L 6775 hCoV-19/Singapore/1054/2020|EPI_ISL_574539|2020-03 ···················· ···················· ····················· ···················· ···················· ····················
```

| | | | | | | | | |
|---|---|---|---|---|---|---|---|---|
| L | 6776 hCoV-19/Singapore/1055/2020\|EPI_ISL_574540\|2020-03 | ···················· | ···················· | ···················· | ···················· | ···················· | ···················· | |
| L | 6777 hCoV-19/USA/SC-DHEC-1134/2020\|EPI_ISL_574560\|2020- | ···················· | ···················· | ···················· | ···················· | ···················· | ···················· | |
| L | 6778 hCoV-19/USA/SC-DHEC-1142/2020\|EPI_ISL_574563\|2020- | ···················· | ···················· | ···················· | ···················· | ···················· | ···················· | |
| L | 6779 hCoV-19/USA/SC-DHEC-1143/2020\|EPI_ISL_574564\|2020- | ···················· | ···················· | ···················· | ···················· | ···················· | ···················· | |
| L | 6780 hCoV-19/USA/SC-DHEC-1144/2020\|EPI_ISL_574565\|2020- | ···················· | ···················· | ···················· | ···················· | ···················· | ···················· | |
| L | 6781 hCoV-19/USA/SC-DHEC-1108/2020\|EPI_ISL_574566\|2020- | ···················· | ···················· | ···················· | ···················· | ···················· | ···················· | |
| L | 6782 hCoV-19/USA/SC-DHEC-1109/2020\|EPI_ISL_574567\|2020- | ···················· | ···················· | ···················· | ···················· | ···················· | ···················· | |
| L | 6783 hCoV-19/USA/SC-DHEC-1113/2020\|EPI_ISL_574568\|2020- | ···················· | ···················· | ···················· | ···················· | ···················· | ···················· | |
| L | 6784 hCoV-19/USA/SC-DHEC-1115/2020\|EPI_ISL_574569\|2020- | ···················· | ···················· | ···················· | ···················· | ···················· | ···················· | |
| L | 6785 hCoV-19/USA/SC-DHEC-1116/2020\|EPI_ISL_574570\|2020- | ···················· | ···················· | ···················· | ···················· | ···················· | ···················· | |
| L | 6786 hCoV-19/USA/SC-DHEC-1117/2020\|EPI_ISL_574571\|2020- | ···················· | ···················· | ···················· | ···················· | ···················· | ···················· | |
| L | 6787 hCoV-19/USA/SC-DHEC-1122/2020\|EPI_ISL_574573\|2020- | ···················· | ···················· | ···················· | ···················· | ···················· | ···················· | |
| L | 6788 hCoV-19/USA/SC-DHEC-1129/2020\|EPI_ISL_574574\|2020- | ···················· | ···················· | ···················· | ···················· | ···················· | ···················· | |
| L | 6789 hCoV-19/USA/SC-DHEC-1131/2020\|EPI_ISL_574575\|2020- | ···················· | ···················· | ···················· | ···················· | ···················· | ···················· | |
| L | 6790 hCoV-19/USA/NM-DOH-2020132668/2020\|EPI_ISL_574576\| | ···················· | ···················· | ···················· | ···················· | ···················· | ···················· | |
| L | 6791 hCoV-19/Brazil/SP-445/2020\|EPI_ISL_574577\|2020-04- | ···················· | ···················· | ···················· | ···················· | ···················· | ···················· | |
| L | 6792 hCoV-19/Brazil/SP-470/2020\|EPI_ISL_574578\|2020-04- | ···················· | ···················· | ···················· | ···················· | ···················· | ···················· | |
| L | 6793 hCoV-19/Brazil/SP-476/2020\|EPI_ISL_574579\|2020-04- | ···················· | ···················· | ···················· | ···················· | ···················· | ···················· | |
| L | 6794 hCoV-19/Brazil/SP-481/2020\|EPI_ISL_574580\|2020-04- | ···················· | ···················· | ···················· | ···················· | ···················· | ···················· | |
| L | 6795 hCoV-19/Brazil/SP-483/2020\|EPI_ISL_574582\|2020-04- | ···················· | ···················· | ···················· | ···················· | ···················· | ···················· | |
| L | 6796 hCoV-19/Brazil/SP-486/2020\|EPI_ISL_574583\|2020-04- | ···················· | ···················· | ···················· | ···················· | ···················· | ···················· | |
| L | 6797 hCoV-19/USA/DE-DHSS-F918831/2020\|EPI_ISL_574585\|20 | ···················· | ···················· | ···················· | ···················· | ···················· | ···················· | |
| L | 6798 hCoV-19/USA/DE-DHSS-F917680/2020\|EPI_ISL_574586\|20 | ···················· | ···················· | ···················· | ···················· | ···················· | ···················· | |
| L | 6799 hCoV-19/Brazil/SP-492/2020\|EPI_ISL_574588\|2020-04- | ···················· | ···················· | ···················· | ···················· | ···················· | ···················· | |
| L | 6800 hCoV-19/Brazil/SP-494/2020\|EPI_ISL_574589\|2020-04- | ···················· | ···················· | ···················· | ···················· | ···················· | ···················· | |
| L | 6801 hCoV-19/Brazil/SP-496/2020\|EPI_ISL_574590\|2020-04- | ···················· | ···················· | ···················· | ···················· | ···················· | ···················· | |
| L | 6802 hCoV-19/Brazil/SP-559/2020\|EPI_ISL_574591\|2020-04- | ···················· | ···················· | ···················· | ···················· | ···················· | ···················· | |
| L | 6803 hCoV-19/Brazil/SP-560/2020\|EPI_ISL_574592\|2020-03- | ···················· | ···················· | ···················· | ···················· | ···················· | ···················· | |
| L | 6804 hCoV-19/Brazil/SP-564/2020\|EPI_ISL_574593\|2020-04- | ···················· | ···················· | ···················· | ···················· | ···················· | ···················· | |
| L | 6805 hCoV-19/Brazil/SP-425/2020\|EPI_ISL_574594\|2020-07- | ···················· | ···················· | ···················· | ···················· | ···················· | ···················· | |
| L | 6806 hCoV-19/Brazil/SP-444/2020\|EPI_ISL_574595\|2020-03- | ···················· | ···················· | ···················· | ···················· | ···················· | ···················· | |
| L | 6807 hCoV-19/Brazil/SP-388/2020\|EPI_ISL_574596\|2020-06- | ···················· | ···················· | ···················· | ···················· | ···················· | ···················· | |
| L | 6808 hCoV-19/Brazil/SP-389/2020\|EPI_ISL_574597\|2020-06- | ···················· | ···················· | ···················· | ···················· | ···················· | ···················· | |
| L | 6809 hCoV-19/Brazil/SP-391/2020\|EPI_ISL_574598\|2020-06- | ···················· | ···················· | ···················· | ···················· | ···················· | ···················· | |
| L | 6810 hCoV-19/USA/DE-DHSS-F918613/2020\|EPI_ISL_574599\|20 | ···················· | ···················· | ···················· | ···················· | ···················· | ···················· | |
| L | 6811 hCoV-19/England/MILK-9748D9/2020\|EPI_ISL_575810\|20 | ···················· | ···················· | ···················· | ···················· | ···················· | ···················· | |
| L | 6812 hCoV-19/Wales/MILK-973148/2020\|EPI_ISL_575811\|2020 | ···················· | ···················· | ···················· | ···················· | ···················· | ···················· | |
| L | 6813 hCoV-19/England/MILK-9730B4/2020\|EPI_ISL_575813\|20 | ···················· | ···················· | ···················· | ···················· | ···················· | ···················· | |
| L | 6814 hCoV-19/England/MILK-974851/2020\|EPI_ISL_575814\|20 | ···················· | ···················· | ···················· | ···················· | ···················· | ···················· | |
| L | 6815 hCoV-19/England/MILK-976576/2020\|EPI_ISL_575815\|20 | ···················· | ···················· | ···················· | ···················· | ···················· | ···················· | |
| L | 6816 hCoV-19/England/MILK-974569/2020\|EPI_ISL_575816\|20 | ···················· | ·A·················· | ···················· | ···················· | ···················· | ···················· | |
| L | 6817 hCoV-19/England/MILK-97489D/2020\|EPI_ISL_575817\|20 | ···················· | ···················· | ···················· | ···················· | ···················· | ···················· | |
| L | 6818 hCoV-19/England/MILK-9764A6/2020\|EPI_ISL_575818\|20 | ···················· | ···················· | ···················· | ···················· | ···················· | ···················· | |
| L | 6819 hCoV-19/England/MILK-973245/2020\|EPI_ISL_575819\|20 | ···················· | ···················· | ···················· | ···················· | ···················· | ···················· | |
| L | 6820 hCoV-19/England/MILK-9730C3/2020\|EPI_ISL_575820\|20 | ···················· | ···················· | ···················· | ···················· | ···················· | ···················· | |
| L | 6821 hCoV-19/Wales/MILK-9762CA/2020\|EPI_ISL_575823\|2020 | ···················· | ···················· | ···················· | ···················· | ···················· | ···················· | |
| L | 6822 hCoV-19/England/MILK-9748F7/2020\|EPI_ISL_575824\|20 | ···················· | ···················· | ···················· | ···················· | ···················· | ···················· | |
| L | 6823 hCoV-19/England/MILK-9746DF/2020\|EPI_ISL_575825\|20 | ···················· | ···················· | ···················· | ···················· | ···················· | ···················· | |
| L | 6824 hCoV-19/England/MILK-976637/2020\|EPI_ISL_575826\|20 | ···················· | ···················· | ···················· | ···················· | ···················· | ···················· | |
| L | 6825 hCoV-19/England/MILK-9766FB/2020\|EPI_ISL_575827\|20 | ···················· | ···················· | ···················· | ···················· | ···················· | ···················· | |
| L | 6826 hCoV-19/England/MILK-9748BB/2020\|EPI_ISL_575828\|20 | ···················· | ···················· | ···················· | ···················· | ···················· | ···················· | |
| L | 6827 hCoV-19/England/MILK-9749C7/2020\|EPI_ISL_575829\|20 | ···················· | ···················· | ···················· | ···················· | ···················· | ···················· | |
| L | 6828 hCoV-19/England/MILK-9735E2/2020\|EPI_ISL_575830\|20 | ···················· | ···················· | ···················· | ···················· | ···················· | ···················· | |
| L | 6829 hCoV-19/England/MILK-9732FA/2020\|EPI_ISL_575831\|20 | ···················· | ···················· | ···················· | ···················· | ···················· | ···················· | |
| L | 6830 hCoV-19/England/MILK-97337F/2020\|EPI_ISL_575832\|20 | ···················· | ···················· | ···················· | ···················· | ···················· | ···················· | |
| L | 6831 hCoV-19/England/MILK-976585/2020\|EPI_ISL_575833\|20 | ···················· | ···················· | ···················· | ···················· | ···················· | ···················· | |
| L | 6832 hCoV-19/England/MILK-9762AC/2020\|EPI_ISL_575834\|20 | ···················· | ···················· | ···················· | ···················· | ···················· | ···················· | |
| L | 6833 hCoV-19/England/MILK-973403/2020\|EPI_ISL_575835\|20 | ···················· | ···················· | ···················· | ···················· | ···················· | ···················· | |
| L | 6834 hCoV-19/England/MILK-97679E/2020\|EPI_ISL_575836\|20 | ···················· | ···················· | ···················· | ···················· | ···················· | ···················· | |
| L | 6835 hCoV-19/England/MILK-9735B5/2020\|EPI_ISL_575838\|20 | ···················· | ···················· | ···················· | ···················· | ···················· | ···················· | |
| L | 6836 hCoV-19/England/MILK-973676/2020\|EPI_ISL_575839\|20 | ···················· | ···················· | ···················· | ···················· | ···················· | ···················· | |
| L | 6837 hCoV-19/England/MILK-976691/2020\|EPI_ISL_575840\|20 | ···················· | ···················· | ···················· | ···················· | ···················· | ···················· | |
| L | 6838 hCoV-19/England/MILK-9765B2/2020\|EPI_ISL_575841\|20 | ··················T· | ···················· | ···················· | ···················· | ···················· | ···················· | 1 |
| L | 6839 hCoV-19/Wales/MILK-973175/2020\|EPI_ISL_575842\|2020 | ···················· | ···················· | ···················· | ···················· | ···················· | ···················· | |
| L | 6840 hCoV-19/England/MILK-976804/2020\|EPI_ISL_575843\|20 | ···················· | ···················· | ···················· | ···················· | ···················· | ···················· | |
| L | 6841 hCoV-19/England/MILK-9730D2/2020\|EPI_ISL_575846\|20 | ···················· | ···················· | ···················· | ···················· | ···················· | ···················· | |
| L | 6842 hCoV-19/England/MILK-9766A0/2020\|EPI_ISL_575847\|20 | ···················· | ···················· | ···················· | ···················· | ···················· | ···················· | |
| L | 6843 hCoV-19/England/MILK-9745D2/2020\|EPI_ISL_575848\|20 | ···················· | ···················· | ···················· | ···················· | ···················· | ···················· | |
| L | 6844 hCoV-19/England/MILK-9749B8/2020\|EPI_ISL_575849\|20 | ···················· | ···················· | ···················· | ···················· | ······T············· | ···················· | |
| L | 6845 hCoV-19/England/MILK-973649/2020\|EPI_ISL_575851\|20 | ········T··········· | ···················· | ···················· | ···················· | ···················· | ···················· | |
| L | 6846 hCoV-19/England/MILK-9732EB/2020\|EPI_ISL_575852\|20 | ···················· | ···················· | ···················· | ···················· | ···················· | ···················· | |
| L | 6847 hCoV-19/England/MILK-9767CB/2020\|EPI_ISL_575853\|20 | ···················· | ···················· | ···················· | ···················· | ···················· | ···················· | |
| L | 6848 hCoV-19/England/MILK-9733F7/2020\|EPI_ISL_575854\|20 | ···················· | ···················· | ···················· | ···················· | ···················· | ···················· | |
| L | 6849 hCoV-19/England/MILK-9731C0/2020\|EPI_ISL_575855\|20 | ···················· | ···················· | ···················· | ···················· | ···················· | ···················· | |
| L | 6850 hCoV-19/Wales/MILK-97351F/2020\|EPI_ISL_575857\|2020 | ···················· | ···················· | ···················· | ···················· | ···················· | ···················· | |
| L | 6851 hCoV-19/England/MILK-97454B/2020\|EPI_ISL_575858\|20 | ···················· | ···················· | ···················· | ···················· | ···················· | ···················· | |
| L | 6852 hCoV-19/England/MILK-976707/2020\|EPI_ISL_575859\|20 | ···················· | ·········A·········· | ···················· | ···················· | ······T············· | ···················· | |
| L | 6853 hCoV-19/England/MILK-976330/2020\|EPI_ISL_575860\|20 | ···················· | ···················· | ···················· | ···················· | ···················· | ···················· | |
| L | 6854 hCoV-19/England/MILK-9747FA/2020\|EPI_ISL_575861\|20 | ···················· | ···················· | ···················· | ···················· | ···················· | ···················· | |
| L | 6855 hCoV-19/England/MILK-974763/2020\|EPI_ISL_575862\|20 | ···················· | ···················· | ···················· | ···················· | ···················· | ···················· | |

| | | | | | | | |
|---|---|---|---|---|---|---|---|
| L | 6856 hCoV-19/England/MILK-976822/2020\|EPI_ISL_575863\|20 | ···················· | ···················· | ····················· | ···················· | ···················· | ···················· |
| L | 6857 hCoV-19/England/MILK-974587/2020\|EPI_ISL_575864\|20 | ···················· | ···················· | ····················· | ···················· | ···················· | ···················· |
| L | 6858 hCoV-19/England/MILK-973281/2020\|EPI_ISL_575865\|20 | ···················· | ···················· | ···········T········· | ···················· | ···················· | ···················· |
| L | 6859 hCoV-19/England/MILK-9744D5/2020\|EPI_ISL_575866\|20 | ···················· | ···················· | ····················· | ···················· | ···················· | ···················· |
| L | 6860 hCoV-19/Wales/MILK-973157/2020\|EPI_ISL_575867\|2020 | ···················· | ···················· | ····················· | ···················· | ···················· | ···················· |
| L | 6861 hCoV-19/England/MILK-9763D6/2020\|EPI_ISL_575868\|20 | ···················· | ···················· | ····················· | ···················· | ···················· | ···················· |
| L | 6862 hCoV-19/England/MILK-976549/2020\|EPI_ISL_575869\|20 | ···················· | ···················· | ····················· | ···················· | ···················· | ···················· |
| L | 6863 hCoV-19/England/MILK-9732BE/2020\|EPI_ISL_575872\|20 | ···················· | ···················· | ····················· | ···················· | ···················· | ···················· |
| L | 6864 hCoV-19/England/MILK-97455A/2020\|EPI_ISL_575873\|20 | ···················· | ···················· | ····················· | ···················· | ···················· | ···················· |
| L | 6865 hCoV-19/England/MILK-9746C0/2020\|EPI_ISL_575890\|20 | ···················· | ···················· | ····················· | ···················· | ···················· | ···················· |
| L | 6866 hCoV-19/England/MILK-976655/2020\|EPI_ISL_575891\|20 | ···················· | ···················· | ····················· | ···················· | ···················· | ···················· |
| L | 6867 hCoV-19/England/MILK-974578/2020\|EPI_ISL_575894\|20 | ···················· | ···················· | ····················· | ···················· | ···················· | ···················· |
| L | 6868 hCoV-19/England/MILK-976479/2020\|EPI_ISL_575896\|20 | ···················· | ···················· | ····················· | ···················· | ···················· | ···················· |
| L | 6869 hCoV-19/England/MILK-976664/2020\|EPI_ISL_575897\|20 | ···················· | ···················· | ····················· | ···················· | ···················· | ···················· |
| L | 6870 hCoV-19/England/MILK-976242/2020\|EPI_ISL_575898\|20 | ···················· | ···················· | ····················· | ···················· | ···················· | ···················· |
| L | 6871 hCoV-19/England/MILK-97345E/2020\|EPI_ISL_575899\|20 | ···················· | ···················· | ····················· | ···················· | ···················· | ···················· |
| L | 6872 hCoV-19/USA/MI-UM-10033759721/2020\|EPI_ISL_578090\| | ···················· | ···················· | ····················· | ···················· | ···················· | ···················· |
| L | 6875 hCoV-19/USA/MI-UM-10033818339/2020\|EPI_ISL_578094\| | ···················· | ···················· | ····················· | ···················· | ···················· | ···················· |
| L | 6876 hCoV-19/USA/MI-UM-10033759038/2020\|EPI_ISL_578095\| | ···················· | ···················· | ····················· | ···················· | ···················· | ···················· |
| L | 6877 hCoV-19/USA/MI-UM-10033760952/2020\|EPI_ISL_578096\| | ···················· | ···················· | ····················· | ···················· | ···················· | ···················· |
| L | 6878 hCoV-19/USA/MI-UM-10033766580/2020\|EPI_ISL_578097\| | ···················· | ···················· | ····················· | ···················· | ···················· | ···················· |
| L | 6879 hCoV-19/USA/MI-UM-10033767074/2020\|EPI_ISL_578098\| | ···················· | ···················· | ····················· | ···················· | ···················· | ···················· |
| L | 6880 hCoV-19/USA/MI-UM-10033768049/2020\|EPI_ISL_578099\| | ···················· | ···················· | ····················· | ···················· | ···················· | ···················· |
| L | 6885 hCoV-19/New Zealand/20VR2662/2020\|EPI_ISL_579326\|2 | ···················· | ···················· | ····················· | ···················· | ···················· | ···················· |
| L | 6886 hCoV-19/New Zealand/20VR2666/2020\|EPI_ISL_579328\|2 | ···················· | ···················· | ····················· | ···················· | ···················· | ···················· |
| L | 6888 hCoV-19/Belgium/ULG-10436/2020\|EPI_ISL_581614\|2020 | ···················· | ···················· | ····················· | ···················· | ···················· | ···················· |
| L | 6889 hCoV-19/Belgium/ULG-10437/2020\|EPI_ISL_581615\|2020 | ···················· | ···················· | ····················· | ···················· | ···················· | ···················· |
| L | 6890 hCoV-19/Belgium/ULG-10438/2020\|EPI_ISL_581616\|2020 | ···················· | ····A··············· | ····················· | ···················· | ···················· | ···················· |
| L | 6891 hCoV-19/Belgium/ULG-10443/2020\|EPI_ISL_581620\|2020 | ···················· | ···················· | ····················· | ···················· | ···················· | ···················· |
| L | 6892 hCoV-19/Belgium/ULG-10444/2020\|EPI_ISL_581621\|2020 | ···················· | ···················· | ····················· | ···················· | ···················· | ···················· |
| L | 6893 hCoV-19/Belgium/ULG-10446/2020\|EPI_ISL_581622\|2020 | ···················· | ···················· | ····················· | ···················· | ···················· | ···················· |
| L | 6894 hCoV-19/Belgium/ULG-10447/2020\|EPI_ISL_581623\|2020 | ···················· | ···················· | ····················· | ···················· | ···················· | ···················· |
| L | 6895 hCoV-19/Belgium/ULG-10449/2020\|EPI_ISL_581624\|2020 | ···················· | ·A·················· | ····················· | ···················· | ···················· | ···················· |
| L | 6898 hCoV-19/Belgium/ULG-10453/2020\|EPI_ISL_581627\|2020 | ···················· | ···················· | ····················· | ···················· | ···················· | ···················· |
| L | 6900 hCoV-19/Belgium/ULG-10455/2020\|EPI_ISL_581629\|2020 | ···················· | ···················· | ····················· | ···················· | ···················· | ···················· |
| L | 6901 hCoV-19/USA/CA-CZB-7204/2020\|EPI_ISL_582940\|2020-0 | ···················· | ···················· | ····················· | ···················· | ···················· | ···················· |
| L | 6902 hCoV-19/USA/CA-CZB-7205/2020\|EPI_ISL_582941\|2020-0 | ···················· | ···················· | ····················· | ···················· | ···················· | ···················· |
| L | 6903 hCoV-19/USA/CA-CZB-7207/2020\|EPI_ISL_582942\|2020-0 | ···················· | ·A·················· | ····················· | ···················· | ···················· | ···················· |
| L | 6904 hCoV-19/USA/CA-CZB-11008/2020\|EPI_ISL_582950\|2020- | ···················· | ···················· | ····················· | ···················· | ···················· | ···················· |
| L | 6905 hCoV-19/USA/CA-CZB-11009/2020\|EPI_ISL_582951\|2020- | ···················· | ·A·················· | ····················· | ···················· | ···················· | ···················· |
| L | 6906 hCoV-19/USA/CA-CZB-11010/2020\|EPI_ISL_582952\|2020- | ···················· | ···················· | ····················· | ···················· | ···················· | ···················· |
| L | 6907 hCoV-19/USA/CA-CZB-11011/2020\|EPI_ISL_582953\|2020- | ···················· | ···················· | ····················· | ···················· | ···················· | ···················· |
| L | 6908 hCoV-19/USA/CA-CZB-11012/2020\|EPI_ISL_582954\|2020- | ···················· | ·A·················· | ····················· | ···················· | ···················· | ···················· |
| L | 6909 hCoV-19/USA/CA-CZB-11015/2020\|EPI_ISL_582955\|2020- | ···················· | ···················· | ····················· | ···················· | ···················· | ···················· |
| L | 6910 hCoV-19/USA/CA-CZB-11017/2020\|EPI_ISL_582957\|2020- | ···················· | ···················· | ····················· | ···················· | ···················· | ···················· |
| L | 6911 hCoV-19/USA/CA-CZB-11018/2020\|EPI_ISL_582958\|2020- | ···················· | ·A·················· | ····················· | ···················· | ···················· | ···················· |
| L | 6912 hCoV-19/USA/CA-CZB-5834/2020\|EPI_ISL_582959\|2020-0 | ···················· | ·A·················· | ····················· | ···················· | ···················· | ···················· |
| L | 6913 hCoV-19/USA/ID-QDX-605/2020\|EPI_ISL_571300\|2020-03 | ···················· | ···················· | ····················· | ···················· | ···················· | ···················· |
| L | 6914 hCoV-19/USA/ID-QDX-606/2020\|EPI_ISL_571301\|2020-03 | ···················· | ···················· | ····················· | ···················· | ···················· | ···················· |
| L | 6915 hCoV-19/USA/ID-QDX-607/2020\|EPI_ISL_571302\|2020-03 | ···················· | ···················· | ····················· | ···················· | ···················· | ···················· |
| L | 6916 hCoV-19/USA/ID-QDX-609/2020\|EPI_ISL_571303\|2020-03 | ···················· | ···················· | ····················· | ···················· | ···················· | ···················· |
| L | 6917 hCoV-19/USA/ID-QDX-610/2020\|EPI_ISL_571304\|2020-03 | ···················· | ···················· | ····················· | ···················· | ···················· | ···················· |
| L | 6918 hCoV-19/USA/ID-QDX-631/2020\|EPI_ISL_571305\|2020-03 | ···················· | ···················· | ····················· | ···················· | ···················· | ···················· |
| L | 6919 hCoV-19/USA/ID-QDX-650/2020\|EPI_ISL_571306\|2020-03 | ···················· | ···················· | ····················· | ···················· | ···················· | ···················· |
| L | 6920 hCoV-19/USA/KS-QDX-523/2020\|EPI_ISL_571307\|2020-03 | ···················· | ···················· | ····················· | ···················· | ···················· | ···················· |
| L | 6921 hCoV-19/USA/LA-QDX-584/2020\|EPI_ISL_571308\|2020-03 | ···················· | ···················· | ····················· | ···················· | ···················· | ···················· |
| L | 6922 hCoV-19/USA/MA-QDX-525/2020\|EPI_ISL_571309\|2020-03 | ···················· | ···················· | ····················· | ···················· | ···················· | ···················· |
| L | 6923 hCoV-19/USA/ME-QDX-633/2020\|EPI_ISL_571310\|2020-03 | ···················· | ···················· | ····················· | ···················· | ···················· | ···················· |
| L | 6924 hCoV-19/USA/ME-QDX-649/2020\|EPI_ISL_571311\|2020-03 | ···················· | ···················· | ····················· | ···················· | ···················· | ···················· |
| L | 6925 hCoV-19/USA/MI-QDX-517/2020\|EPI_ISL_571312\|2020-03 | ···················· | ···················· | ····················· | ···················· | ···················· | ···················· |
| L | 6926 hCoV-19/USA/MI-QDX-555/2020\|EPI_ISL_571313\|2020-03 | ···················· | ···················· | ····················· | ···················· | ···················· | ···················· |
| L | 6927 hCoV-19/USA/MI-QDX-558/2020\|EPI_ISL_571314\|2020-03 | ···················· | ···················· | ····················· | ···················· | ···················· | ···················· |
| L | 6928 hCoV-19/USA/MI-QDX-559/2020\|EPI_ISL_571315\|2020-03 | ···················· | ···················· | ····················· | ···················· | ···················· | ···················· |
| L | 6929 hCoV-19/USA/MI-QDX-560/2020\|EPI_ISL_571316\|2020-03 | ···················· | ···················· | ····················· | ···················· | ···················· | ···················· |
| L | 6930 hCoV-19/USA/MI-QDX-561/2020\|EPI_ISL_571317\|2020-03 | ···················· | ···················· | ····················· | ···················· | ···················· | ···················· |
| L | 6931 hCoV-19/USA/MI-QDX-562/2020\|EPI_ISL_571318\|2020-03 | ···················· | ···················· | ····················· | ···················· | ···················· | ···················· |
| L | 6932 hCoV-19/USA/MI-QDX-565/2020\|EPI_ISL_571319\|2020-03 | ···················· | ···················· | ····················· | ···················· | ···················· | ···················· |
| L | 6933 hCoV-19/USA/MI-QDX-567/2020\|EPI_ISL_571320\|2020-03 | ···················· | ···················· | ····················· | ···················· | ···················· | ···················· |
| L | 6934 hCoV-19/USA/MI-QDX-568/2020\|EPI_ISL_571321\|2020-03 | ···················· | ···················· | ····················· | ···················· | ···················· | ···················· |
| L | 6935 hCoV-19/USA/MI-QDX-569/2020\|EPI_ISL_571322\|2020-03 | ···················· | ···················· | ····················· | ···················· | ···················· | ···················· |
| L | 6936 hCoV-19/USA/MI-QDX-571/2020\|EPI_ISL_571323\|2020-03 | ···················· | ···················· | ····················· | ···················· | ···················· | ···················· |
| L | 6937 hCoV-19/USA/MI-QDX-576/2020\|EPI_ISL_571324\|2020-03 | ···················· | ···················· | ····················· | ···················· | ···················· | ···················· |
| L | 6938 hCoV-19/USA/MI-QDX-587/2020\|EPI_ISL_571325\|2020-03 | ···················· | ···················· | ····················· | ···················· | ···················· | ···················· |
| L | 6939 hCoV-19/USA/MI-QDX-588/2020\|EPI_ISL_571326\|2020-03 | ···················· | ···················· | ····················· | ···················· | ···················· | ···················· |
| L | 6940 hCoV-19/USA/MI-QDX-589/2020\|EPI_ISL_571327\|2020-03 | ···················· | ···················· | ····················· | ···················· | ···················· | ···················· |
| L | 6941 hCoV-19/USA/MI-QDX-590/2020\|EPI_ISL_571328\|2020-03 | ···················· | ···················· | ····················· | ···················· | ···················· | ···················· |
| L | 6942 hCoV-19/USA/MI-QDX-591/2020\|EPI_ISL_571329\|2020-03 | ···················· | ···················· | ····················· | ···················· | ···················· | ···················· |
| L | 6943 hCoV-19/USA/MI-QDX-592/2020\|EPI_ISL_571330\|2020-03 | ···················· | ···················· | ····················· | ···················· | ···················· | ···················· |
| L | 6944 hCoV-19/England/CAMC-9C19CB/2020\|EPI_ISL_573212\|20 | ···················· | ···················· | ····················· | ···················· | ···················· | ···················· |
| L | 6945 hCoV-19/England/CAMC-9C19E9/2020\|EPI_ISL_573213\|20 | ···················· | ···················· | ····················· | ···················· | ···················· | ···················· |

| | | | | | | | | |
|---|---|---|---|---|---|---|---|---|
| L | 6946 hCoV-19/England/CAMC-9C1A04/2020\|EPI_ISL_573214\|20 | ···················· | ···················· | ···················· | ···················· | ···················· | ···················· | |
| L | 6947 hCoV-19/England/CAMC-9C1A22/2020\|EPI_ISL_573215\|20 | ···················· | ···················· | ···················· | ···················· | ···················· | ···················· | |
| L | 6948 hCoV-19/England/CAMC-9C1A31/2020\|EPI_ISL_573216\|20 | ···················· | ···················· | ···················· | ···················· | ···················· | ···················· | |
| L | 6949 hCoV-19/England/CAMC-9C1A5F/2020\|EPI_ISL_573217\|20 | ···················· | ···················· | ···················· | ···················· | ···················· | ···················· | |
| L | 6950 hCoV-19/England/CAMC-9C1AC8/2020\|EPI_ISL_573218\|20 | ···················· | ···················· | ···················· | ···················· | ···················· | ···················· | |
| L | 6951 hCoV-19/England/CAMC-9C1AD7/2020\|EPI_ISL_573219\|20 | ···················· | ···················· | ···················· | ···················· | ···················· | ···················· | |
| L | 6952 hCoV-19/England/CAMC-9C1B01/2020\|EPI_ISL_573220\|20 | ···················· | ···················· | ···················· | ···················· | ···················· | ···················· | |
| L | 6953 hCoV-19/England/CAMC-9C1B3E/2020\|EPI_ISL_573221\|20 | ···················· | ···················· | ···················· | ···················· | ···················· | ···················· | |
| L | 6954 hCoV-19/England/CAMC-9C1B4D/2020\|EPI_ISL_573222\|20 | ···················· | ···················· | ···················· | ···················· | ···················· | ···················· | |
| L | 6955 hCoV-19/England/CAMC-9C1B98/2020\|EPI_ISL_573223\|20 | ···················· | ···················· | ···················· | ···················· | ···················· | ···················· | |
| L | 6956 hCoV-19/England/CAMC-9C1BA7/2020\|EPI_ISL_573224\|20 | ···················· | ·A·················· | ···················· | ···················· | ···················· | ··A················· | |
| L | 6957 hCoV-19/England/CAMC-9C1BB6/2020\|EPI_ISL_573225\|20 | ···················· | ···················· | ···················· | ···················· | ···················· | ···················· | |
| L | 6958 hCoV-19/England/CAMC-9C1C0E/2020\|EPI_ISL_573226\|20 | ···················· | ···················· | ···················· | ···················· | ···················· | ···················· | |
| L | 6959 hCoV-19/England/CAMC-9C1C3B/2020\|EPI_ISL_573227\|20 | ···················· | ···················· | ···················· | ···················· | ···················· | ···················· | |
| L | 6960 hCoV-19/England/CAMC-9C1CC2/2020\|EPI_ISL_573228\|20 | ···················· | ···················· | ···················· | ···················· | ···················· | ···················· | |
| L | 6961 hCoV-19/England/CAMC-9C1CE0/2020\|EPI_ISL_573229\|20 | ···················· | ···················· | ···················· | ···················· | ···················· | ···················· | |
| L | 6962 hCoV-19/England/CAMC-9C1CFF/2020\|EPI_ISL_573230\|20 | ···················· | ···················· | ···················· | ···················· | ···················· | ···················· | |
| L | 6963 hCoV-19/England/CAMC-9C1D29/2020\|EPI_ISL_573231\|20 | ···················· | ···················· | ···················· | ···················· | ···················· | ···················· | |
| L | 6964 hCoV-19/England/CAMC-9C1D83/2020\|EPI_ISL_573233\|20 | ···················· | ···················· | ···················· | ···················· | ···················· | ···················· | |
| L | 6965 hCoV-19/Wales/PHWC-16B873/2020\|EPI_ISL_573910\|2020 | ···················· | ···················· | ···················· | ···················· | ···················· | ···················· | |
| L | 6966 hCoV-19/Wales/PHWC-16B882/2020\|EPI_ISL_573911\|2020 | ···················· | ···················· | ···················· | ···················· | ···················· | ···················· | |
| L | 6967 hCoV-19/Wales/PHWC-16B8A0/2020\|EPI_ISL_573912\|2020 | ···················· | ···················· | ···················· | ···················· | ···················· | ···················· | |
| L | 6968 hCoV-19/Wales/PHWC-16B8BF/2020\|EPI_ISL_573913\|2020 | ···················· | ···················· | ···················· | ···················· | ···················· | ···················· | |
| L | 6969 hCoV-19/Wales/PHWC-16B8DD/2020\|EPI_ISL_573914\|2020 | ···················· | ···················· | ···················· | ···················· | ···················· | ···················· | |
| L | 6970 hCoV-19/Wales/PHWC-16B8EC/2020\|EPI_ISL_573915\|2020 | ···················· | ···················· | ···················· | ···················· | ···················· | ···················· | |
| L | 6972 hCoV-19/Wales/PHWC-16B907/2020\|EPI_ISL_573917\|2020 | ···················· | ···················· | ···················· | ···················· | ···················· | ···················· | |
| L | 6973 hCoV-19/Wales/PHWC-16B925/2020\|EPI_ISL_573918\|2020 | ···················· | ···················· | ···················· | ···················· | ···················· | ···················· | |
| L | 6974 hCoV-19/Wales/PHWC-16B934/2020\|EPI_ISL_573919\|2020 | ···················· | ···················· | ···················· | ···················· | ···················· | ···················· | |
| L | 6975 hCoV-19/Wales/PHWC-16B943/2020\|EPI_ISL_573920\|2020 | ···················· | ···················· | ···················· | ···················· | ···················· | ···················· | |
| L | 6976 hCoV-19/Wales/PHWC-16B952/2020\|EPI_ISL_573921\|2020 | ···················· | ···················· | ···················· | ···················· | ···················· | ···················· | |
| L | 6977 hCoV-19/Wales/PHWC-16B961/2020\|EPI_ISL_573922\|2020 | ···················· | ···················· | ···················· | ···················· | ···················· | ···················· | |
| L | 6978 hCoV-19/Wales/PHWC-16B970/2020\|EPI_ISL_573923\|2020 | ···················· | ···················· | ···················· | ···················· | ···················· | ···················· | |
| L | 6979 hCoV-19/Wales/PHWC-16B99E/2020\|EPI_ISL_573924\|2020 | ···················· | ···················· | ···················· | ···················· | ···················· | ···················· | |
| L | 6980 hCoV-19/Wales/PHWC-16B9AD/2020\|EPI_ISL_573925\|2020 | ···················· | ···················· | ···················· | ···················· | ···················· | ···················· | |
| L | 6981 hCoV-19/Wales/PHWC-16B9CB/2020\|EPI_ISL_573926\|2020 | ···················· | ···················· | ···················· | ···················· | ···················· | ···················· | |
| L | 6982 hCoV-19/Wales/PHWC-16B9E9/2020\|EPI_ISL_573927\|2020 | ···················· | ···················· | ···················· | ···················· | ···················· | ···················· | |
| L | 6984 hCoV-19/Wales/PHWC-16BA04/2020\|EPI_ISL_573929\|2020 | ···················· | ···················· | ···················· | ···················· | ···················· | ···················· | |
| L | 6985 hCoV-19/Wales/PHWC-16BA22/2020\|EPI_ISL_573930\|2020 | ···················· | ···················· | ···················· | ···················· | ···················· | ···················· | |
| L | 6986 hCoV-19/Wales/PHWC-16BA31/2020\|EPI_ISL_573931\|2020 | ···················· | ···················· | ···················· | ···················· | ···················· | ···················· | |
| L | 6987 hCoV-19/Wales/PHWC-16BA40/2020\|EPI_ISL_573932\|2020 | ···················· | ···················· | ···················· | ···················· | ···················· | ···················· | |
| L | 6988 hCoV-19/Wales/PHWC-16BA5F/2020\|EPI_ISL_573933\|2020 | ···················· | ···················· | ···················· | ···················· | ···················· | ···················· | |
| L | 6989 hCoV-19/Wales/PHWC-16BA6E/2020\|EPI_ISL_573934\|2020 | ···················· | ···················· | ···················· | ···················· | ···················· | ···················· | |
| L | 6990 hCoV-19/Wales/PHWC-16BA7D/2020\|EPI_ISL_573935\|2020 | ···················· | ···················· | ···················· | ···················· | ···················· | ···················· | |
| L | 6991 hCoV-19/Wales/PHWC-16BA8C/2020\|EPI_ISL_573936\|2020 | ···················· | ···················· | ···················· | ···················· | ···················· | ···················· | |
| L | 6992 hCoV-19/Wales/PHWC-16BAAA/2020\|EPI_ISL_573937\|2020 | ···················· | ···················· | ···················· | ···················· | ···················· | ···················· | |
| L | 6993 hCoV-19/Wales/PHWC-16BAB9/2020\|EPI_ISL_573938\|2020 | ···················· | ···················· | ···················· | ···················· | ···················· | ···················· | |
| L | 6994 hCoV-19/Wales/PHWC-16BAC8/2020\|EPI_ISL_573939\|2020 | ···················· | ···················· | ···················· | ···················· | ···················· | ···················· | |
| L | 6996 hCoV-19/Wales/PHWC-16BAE6/2020\|EPI_ISL_573941\|2020 | ···················· | ···················· | ···················· | ···················· | ···················· | ···················· | |
| L | 6997 hCoV-19/Wales/PHWC-16BAF5/2020\|EPI_ISL_573942\|2020 | ···················· | ···················· | ···················· | ···················· | ···················· | ···················· | |
| L | 6998 hCoV-19/Wales/PHWC-16BB2F/2020\|EPI_ISL_573943\|2020 | ···················· | ···················· | ···················· | ···················· | NNN················· | ···················· | 1 |
| L | 6999 hCoV-19/Wales/PHWC-16BB3E/2020\|EPI_ISL_573944\|2020 | ···················· | ···················· | ···················· | ···················· | ···················· | ···················· | |
| L | 7000 hCoV-19/Wales/PHWC-16BB4D/2020\|EPI_ISL_573945\|2020 | ···················· | ···················· | ···················· | ···················· | ···················· | ···················· | |
| L | 7001 hCoV-19/Wales/PHWC-16BB5C/2020\|EPI_ISL_573946\|2020 | ···················· | ···················· | ···················· | ···················· | ···················· | ···················· | |
| L | 7003 hCoV-19/Wales/PHWC-16BB7A/2020\|EPI_ISL_573948\|2020 | ···················· | ···················· | ···················· | ···················· | ···················· | ···················· | |
| L | 7004 hCoV-19/Wales/PHWC-16BB89/2020\|EPI_ISL_573949\|2020 | ···················· | ···················· | ···················· | ···················· | ···················· | ···················· | |
| L | 7005 hCoV-19/Wales/PHWC-16BB98/2020\|EPI_ISL_573950\|2020 | ···················· | ···················· | ···················· | ···················· | ···················· | ···················· | |
| L | 7006 hCoV-19/Wales/PHWC-16BBB6/2020\|EPI_ISL_573951\|2020 | ···················· | ···················· | ···················· | ···················· | ···················· | ···················· | |
| L | 7008 hCoV-19/Wales/PHWC-16BBD4/2020\|EPI_ISL_573953\|2020 | ···················· | ···················· | ···················· | ···················· | ···················· | ···················· | |
| L | 7009 hCoV-19/Wales/PHWC-16BC0E/2020\|EPI_ISL_573954\|2020 | ···················· | ···················· | ···················· | ···················· | ···················· | ···················· | |
| L | 7010 hCoV-19/Wales/PHWC-16BC3B/2020\|EPI_ISL_573955\|2020 | ···················· | ···················· | ···················· | ···················· | ···················· | ···················· | |
| L | 7011 hCoV-19/Wales/PHWC-16BC4A/2020\|EPI_ISL_573956\|2020 | ···················· | ···················· | ···················· | ···················· | ···················· | ···················· | |
| L | 7012 hCoV-19/Wales/PHWC-16BC59/2020\|EPI_ISL_573957\|2020 | ···················· | ···················· | ···················· | ···················· | ···················· | ···················· | |
| L | 7013 hCoV-19/Wales/PHWC-16BC68/2020\|EPI_ISL_573958\|2020 | ···················· | ···················· | ···················· | ···················· | ···················· | ···················· | |
| L | 7014 hCoV-19/Wales/PHWC-16BC77/2020\|EPI_ISL_573959\|2020 | ···················· | ···················· | ···················· | ···················· | ···················· | ···················· | |
| L | 7015 hCoV-19/Wales/PHWC-16BC95/2020\|EPI_ISL_573960\|2020 | ···················· | ···················· | ···················· | ···················· | ···················· | ···················· | |
| L | 7016 hCoV-19/Wales/PHWC-16BCA4/2020\|EPI_ISL_573961\|2020 | ···················· | ···················· | ···················· | ···················· | ···················· | ···················· | |
| L | 7017 hCoV-19/Wales/PHWC-16BCB3/2020\|EPI_ISL_573962\|2020 | ···················· | ···················· | ···················· | ···················· | ···················· | ···················· | |
| L | 7018 hCoV-19/Wales/PHWC-16BCC2/2020\|EPI_ISL_573963\|2020 | ···················· | ···················· | ···················· | ···················· | ···················· | ···················· | |
| L | 7019 hCoV-19/Wales/PHWC-16BCD1/2020\|EPI_ISL_573964\|2020 | ···················· | ···················· | ···················· | ···················· | ···················· | ···················· | |
| L | 7020 hCoV-19/Wales/PHWC-16BCFF/2020\|EPI_ISL_573965\|2020 | ···················· | ···················· | ···················· | ···················· | ···················· | ···················· | |
| L | 7021 hCoV-19/Wales/PHWC-16BD29/2020\|EPI_ISL_573966\|2020 | ···················· | ···················· | ···················· | ···················· | ···················· | ···················· | |
| L | 7022 hCoV-19/Wales/PHWC-16BD38/2020\|EPI_ISL_573967\|2020 | ···················· | ···················· | ···················· | ···················· | ···················· | ···················· | |
| L | 7024 hCoV-19/Wales/PHWC-16BD65/2020\|EPI_ISL_573969\|2020 | ···················· | ···················· | ···················· | ···················· | ···················· | ···················· | |
| L | 7025 hCoV-19/Wales/PHWC-16BD74/2020\|EPI_ISL_573970\|2020 | ···················· | ···················· | ···················· | ···················· | ······T············· | ···················· | |
| L | 7026 hCoV-19/Wales/PHWC-16BD92/2020\|EPI_ISL_573971\|2020 | ···················· | ···················· | ···················· | ···················· | ···················· | ···················· | |
| L | 7027 hCoV-19/Wales/PHWC-16BDA1/2020\|EPI_ISL_573972\|2020 | ···················· | ···················· | ···················· | ···················· | ···················· | ···················· | |
| L | 7028 hCoV-19/Wales/PHWC-16BDB0/2020\|EPI_ISL_573973\|2020 | ···················· | ···················· | ···················· | ···················· | ···················· | ···················· | |
| L | 7029 hCoV-19/Wales/PHWC-16BDDE/2020\|EPI_ISL_573974\|2020 | ···················· | ···················· | ···················· | ···················· | ···················· | ···················· | |
| L | 7030 hCoV-19/Wales/PHWC-16BDED/2020\|EPI_ISL_573975\|2020 | ···················· | ···················· | ···················· | ···················· | ···················· | ···················· | |
| L | 7031 hCoV-19/Wales/PHWC-16BDFC/2020\|EPI_ISL_573976\|2020 | ···················· | ···················· | ···················· | ···················· | ···················· | ···················· | |

| | | | | | | | | |
|---|---|---|---|---|---|---|---|---|
| L | 7032 | hCoV-19/Wales/PHWC-16BE08/2020\|EPI_ISL_573977\|2020 | ···················· | ···················· | ···················· | ···················· | ···················· | ···················· |
| L | 7033 | hCoV-19/Wales/PHWC-16BE17/2020\|EPI_ISL_573978\|2020 | ···················· | ···················· | ···················· | ···················· | ···················· | ···················· |
| L | 7034 | hCoV-19/Wales/PHWC-16BE35/2020\|EPI_ISL_573979\|2020 | ···················· | ···················· | ···················· | ···················· | ···················· | ···················· |
| L | 7035 | hCoV-19/Wales/PHWC-16BE44/2020\|EPI_ISL_573980\|2020 | ···················· | ···················· | ···················· | ···················· | ···················· | ···················· |
| L | 7036 | hCoV-19/Wales/PHWC-16BE53/2020\|EPI_ISL_573981\|2020 | ···················· | ···················· | ···················· | ···················· | ···················· | ···················· |
| L | 7037 | hCoV-19/Wales/PHWC-16BE62/2020\|EPI_ISL_573982\|2020 | ···················· | ···················· | ···················· | ···················· | ···················· | ···················· |
| L | 7038 | hCoV-19/Wales/PHWC-16BE71/2020\|EPI_ISL_573983\|2020 | ···················· | ···················· | ···················· | ···················· | ···················· | ···················· |
| L | 7039 | hCoV-19/Wales/PHWC-16BE80/2020\|EPI_ISL_573984\|2020 | ···················· | ···················· | ···················· | ···················· | ···················· | ···················· |
| L | 7040 | hCoV-19/Wales/PHWC-16BEBD/2020\|EPI_ISL_573985\|2020 | ···················· | ···················· | ···················· | ···················· | ···················· | ···················· |
| L | 7041 | hCoV-19/Wales/PHWC-16BECC/2020\|EPI_ISL_573986\|2020 | ···················· | ···················· | ···················· | ···················· | ···················· | ···················· |
| L | 7042 | hCoV-19/Wales/PHWC-16BEEA/2020\|EPI_ISL_573987\|2020 | ···················· | ···················· | ···················· | ···················· | ···················· | ···················· |
| L | 7043 | hCoV-19/Wales/PHWC-16BF05/2020\|EPI_ISL_573988\|2020 | ···················· | ···················· | ···················· | ···················· | ···················· | ···················· |
| L | 7044 | hCoV-19/Wales/PHWC-16BF14/2020\|EPI_ISL_573989\|2020 | ···················· | ···················· | ···················· | ···················· | ···················· | ···················· |
| L | 7046 | hCoV-19/Wales/PHWC-16BF41/2020\|EPI_ISL_573991\|2020 | ···················· | ···················· | ···················· | ···················· | ···················· | ···················· |
| L | 7048 | hCoV-19/USA/SC-DHEC-1148/2020\|EPI_ISL_574543\|2020- | ···················· | ·A·················· | ···················· | ···················· | ···················· | ···················· |
| L | 7049 | hCoV-19/USA/SC-DHEC-1150/2020\|EPI_ISL_574544\|2020- | ···················· | ·A·················· | ···················· | ···················· | ···················· | ···················· |
| L | 7050 | hCoV-19/USA/SC-DHEC-1153/2020\|EPI_ISL_574545\|2020- | ···················· | ···················· | ···················· | ···················· | ···················· | ···················· |
| L | 7051 | hCoV-19/USA/SC-DHEC-1154/2020\|EPI_ISL_574546\|2020- | ···················· | ·A·················· | ···················· | ···················· | ···················· | ···················· |
| L | 7052 | hCoV-19/USA/SC-DHEC-1177/2020\|EPI_ISL_574547\|2020- | ···················· | ···················· | ···················· | ···················· | ···················· | ···················· |
| L | 7053 | hCoV-19/USA/SC-DHEC-0543/2020\|EPI_ISL_574550\|2020- | ···················· | ···················· | ···················· | ···················· | ···················· | ···················· |
| L | 7054 | hCoV-19/USA/SC-DHEC-0552/2020\|EPI_ISL_574551\|2020- | ···················· | ···················· | ···················· | ···················· | ···················· | ···················· |
| L | 7055 | hCoV-19/USA/SC-DHEC-0553/2020\|EPI_ISL_574552\|2020- | ···················· | ···················· | ···················· | ···················· | ···················· | ···················· |
| L | 7056 | hCoV-19/USA/SC-DHEC-0554/2020\|EPI_ISL_574553\|2020- | ···················· | ···················· | ···················· | ···················· | ···················· | ···················· |
| L | 7057 | hCoV-19/USA/SC-DHEC-1102/2020\|EPI_ISL_574554\|2020- | ···················· | ···················· | ···················· | ···················· | ···················· | ···················· |
| L | 7058 | hCoV-19/USA/SC-DHEC-1103/2020\|EPI_ISL_574555\|2020- | ···················· | ···················· | ···················· | ···················· | ···················· | ···················· |
| L | 7059 | hCoV-19/USA/SC-DHEC-1107/2020\|EPI_ISL_574557\|2020- | ···················· | ···················· | ···················· | ···················· | ···················· | ···················· |
| L | 7060 | hCoV-19/USA/SC-DHEC-1132/2020\|EPI_ISL_574558\|2020- | ···················· | ···················· | ···················· | ···················· | ···················· | ···················· |
| L | 7061 | hCoV-19/USA/SC-DHEC-1133/2020\|EPI_ISL_574559\|2020- | ···················· | ···················· | ···················· | ···················· | ···················· | ···················· |
| L | 7062 | hCoV-19/USA/SC-DHEC-1138/2020\|EPI_ISL_574561\|2020- | ···················· | ···················· | ···················· | ···················· | ···················· | ···················· |
| L | 7063 | hCoV-19/USA/SC-DHEC-1139/2020\|EPI_ISL_574562\|2020- | ···················· | ···················· | ···················· | ···················· | ···················· | ···················· |
| L | 7064 | hCoV-19/England/MILK-97355B/2020\|EPI_ISL_575874\|20 | ···················· | ···················· | ···················· | ···················· | ···················· | ···················· |
| L | 7065 | hCoV-19/England/MILK-97339D/2020\|EPI_ISL_575876\|20 | ···················· | ···················· | ···················· | ···················· | ···················· | ···················· |
| L | 7066 | hCoV-19/England/MILK-9745F0/2020\|EPI_ISL_575878\|20 | ···················· | ···A················ | ···················· | ···················· | ···················· | ···················· |
| L | 7067 | hCoV-19/England/MILK-97638B/2020\|EPI_ISL_575879\|20 | ···················· | ···················· | ···················· | ···················· | ···················· | ···················· |
| L | 7068 | hCoV-19/England/MILK-9731DF/2020\|EPI_ISL_575880\|20 | ···················· | ···················· | ···················· | ···················· | ···················· | ···················· |
| L | 7069 | hCoV-19/England/MILK-97636D/2020\|EPI_ISL_575881\|20 | ···················· | ···················· | ···················· | ···················· | ···················· | ···················· |
| L | 7070 | hCoV-19/England/MILK-976734/2020\|EPI_ISL_575882\|20 | ···················· | ···················· | ···········T········ | ···················· | ···················· | ···················· |
| L | 7071 | hCoV-19/Northern Ireland/MILK-9746B1/2020\|EPI_ISL_ | ···················· | ···················· | ···················· | ···················· | ···················· | ···················· |
| L | 7072 | hCoV-19/Wales/MILK-973184/2020\|EPI_ISL_575885\|2020 | ···················· | ···················· | ···················· | ···················· | ···················· | ···················· |
| L | 7073 | hCoV-19/England/MILK-97642E/2020\|EPI_ISL_575886\|20 | ···················· | ···················· | ···················· | ···················· | ···················· | ···················· |
| L | 7074 | hCoV-19/England/MILK-9765FE/2020\|EPI_ISL_575887\|20 | ···················· | ·A·················· | ···················· | ···················· | ···················· | ···A················ |
| L | 7075 | hCoV-19/England/MILK-9735C4/2020\|EPI_ISL_575888\|20 | ···················· | ···················· | ···················· | ···················· | ···················· | ···················· |
| L | 7076 | hCoV-19/England/MILK-976725/2020\|EPI_ISL_575889\|20 | ···················· | ···················· | ···················· | ···················· | ···················· | ···················· |
| L | 7077 | hCoV-19/England/MILK-97628E/2020\|EPI_ISL_575892\|20 | ···················· | ···················· | ···················· | ···················· | ···················· | ···················· |
| L | 7078 | hCoV-19/USA/AK-PHL384/2020\|EPI_ISL_576111\|2020-06- | ···················· | ···················· | ···················· | ···················· | ···················· | ···················· |
| L | 7079 | hCoV-19/USA/AK-PHL404/2020\|EPI_ISL_576112\|2020-06- | ···················· | ···················· | ···················· | ···················· | ···················· | ···················· |
| L | 7080 | hCoV-19/Indonesia/YO-UGM-10001/2020\|EPI_ISL_576113 | ···················· | ···A················ | ···················· | ···················· | ···················· | ···················· |
| L | 7081 | hCoV-19/Indonesia/YO-UGM-10004/2020\|EPI_ISL_576114 | ···················· | ···················· | ···················· | ···················· | ···················· | ···················· |
| L | 7082 | hCoV-19/Indonesia/YO-UGM-10003/2020\|EPI_ISL_576115 | ···················· | ···A················ | ···················· | ···················· | ···················· | ···················· |
| L | 7083 | hCoV-19/Indonesia/YO-UGM-10002/2020\|EPI_ISL_576116 | ···················· | ···A················ | ···················· | ···················· | ···················· | ···················· |
| L | 7084 | hCoV-19/Malta/BAL-Sliema-1/2020\|EPI_ISL_576117\|202 | ···················· | ·A·················· | ···················· | ···················· | ···················· | ···················· |
| L | 7085 | hCoV-19/Malta/BAL-Sliema-2/2020\|EPI_ISL_576118\|202 | ···················· | ·A·················· | ···················· | ···················· | ···················· | ···················· |
| L | 7086 | hCoV-19/Malta/BAL-Sliema-4/2020\|EPI_ISL_576120\|202 | ···················· | ·A·················· | ···················· | ···················· | ···················· | ···················· |
| L | 7087 | hCoV-19/Malta/BAL-Sliema-5/2020\|EPI_ISL_576121\|202 | ···················· | ·A·················· | ···················· | ···················· | ···················· | ···················· |
| L | 7088 | hCoV-19/Malta/BAL-Sliema-9/2020\|EPI_ISL_576125\|202 | ···················· | ·A·················· | ···················· | ···················· | ···················· | ···················· |
| L | 7089 | hCoV-19/Australia/QLD1054/2020\|EPI_ISL_576140\|2020 | ···················· | ···················· | ···················· | ···················· | ···················· | ···················· |
| L | 7090 | hCoV-19/Australia/QLD1056/2020\|EPI_ISL_576141\|2020 | ···················· | ···················· | ···················· | ···················· | ···················· | ···················· |
| L | 7091 | hCoV-19/Australia/QLD1057/2020\|EPI_ISL_576142\|2020 | ···················· | ···················· | ···················· | ···················· | ···················· | ···················· |
| L | 7092 | hCoV-19/Australia/QLD1058/2020\|EPI_ISL_576143\|2020 | ···················· | ···················· | ···················· | ···················· | ···················· | ···················· |
| L | 7093 | hCoV-19/Australia/QLD1060/2020\|EPI_ISL_576144\|2020 | ···················· | ···················· | ···················· | ···················· | ···················· | ···················· |
| L | 7094 | hCoV-19/Indonesia/YO-UGM-107727/2020\|EPI_ISL_57614 | ···················· | ···················· | ···················· | ···················· | ···················· | ···················· |
| L | 7095 | hCoV-19/Ukraine/Kharkiv-705/2020\|EPI_ISL_576146\|20 | ···················· | ···················· | ···················· | ···················· | ···················· | ···················· |
| L | 7096 | hCoV-19/Ukraine/Ivano-Frankivsk-706/2020\|EPI_ISL_5 | ···················· | ···················· | ···················· | ···················· | ···················· | ···················· |
| L | 7097 | hCoV-19/Ukraine/Kyiv-785/2020\|EPI_ISL_576148\|2020- | ···················· | ···················· | ···················· | ···················· | ···················· | ···················· |
| L | 7098 | hCoV-19/USA/GA-CDC-4459/2020\|EPI_ISL_576190\|2020-0 | ···················· | ···················· | ···················· | ···················· | ···················· | ···················· |
| L | 7099 | hCoV-19/USA/GA-CDC-4466/2020\|EPI_ISL_576191\|2020-0 | ···················· | ···················· | ···················· | ···················· | ···················· | ···················· |
| L | 7100 | hCoV-19/USA/GA-CDC-6364/2020\|EPI_ISL_576192\|2020-0 | ···················· | ···················· | ···················· | ···················· | ···················· | ···················· |
| L | 7101 | hCoV-19/USA/GA-CDC-6365/2020\|EPI_ISL_576193\|2020-0 | ···················· | ···················· | ···················· | ···················· | ···················· | ···················· |
| L | 7102 | hCoV-19/USA/GA-CDC-6367/2020\|EPI_ISL_576194\|2020-0 | ···················· | ···················· | ···················· | ···················· | ···················· | ···················· |
| L | 7103 | hCoV-19/USA/GA-CDC-6368/2020\|EPI_ISL_576195\|2020-0 | ···················· | ···················· | ···················· | ···················· | ···················· | ···················· |
| L | 7104 | hCoV-19/USA/AR-CDC-0871/2020\|EPI_ISL_576196\|2020-0 | ···················· | ···················· | ···················· | ···················· | ···················· | ···················· |
| L | 7105 | hCoV-19/USA/AR-CDC-7907/2020\|EPI_ISL_576197\|2020-0 | ···················· | ···················· | ···················· | ···················· | ···················· | ···················· |
| L | 7106 | hCoV-19/Wales/PHWC-16D70B/2020\|EPI_ISL_577436\|2020 | ···················· | ···················· | ···················· | ···················· | ···················· | ···················· |
| L | 7107 | hCoV-19/Wales/PHWC-16D71A/2020\|EPI_ISL_577437\|2020 | ···················· | ···················· | ···················· | ···················· | ···················· | ···················· |
| L | 7108 | hCoV-19/Wales/PHWC-16D729/2020\|EPI_ISL_577438\|2020 | ···················· | ···················· | ···················· | ···················· | ···················· | ···················· |
| L | 7109 | hCoV-19/Wales/PHWC-16D738/2020\|EPI_ISL_577439\|2020 | ···················· | ···················· | ···················· | ···················· | ···················· | ···················· |
| L | 7110 | hCoV-19/Wales/PHWC-16D747/2020\|EPI_ISL_577440\|2020 | ···················· | ···················· | ···················· | ···················· | ···················· | ···················· |
| L | 7111 | hCoV-19/Wales/PHWC-16D756/2020\|EPI_ISL_577441\|2020 | ···················· | ···················· | ···················· | ···················· | ···················· | ···················· |
| L | 7112 | hCoV-19/Wales/PHWC-16D765/2020\|EPI_ISL_577442\|2020 | ···················· | ···················· | ···················· | ···················· | ···················· | ···················· |
| L | 7113 | hCoV-19/Wales/PHWC-16D774/2020\|EPI_ISL_577443\|2020 | ···················· | ···················· | ···················· | ···················· | ···················· | ···················· |

```
L 7114 hCoV-19/Wales/PHWC-16D783/2020|EPI_ISL_577444|2020 ···················· ···················· ···················· ···················· ···················· ····················
L 7115 hCoV-19/Wales/PHWC-16D792/2020|EPI_ISL_577445|2020 ···················· ···················· ···················· ···················· ···················· ····················
L 7116 hCoV-19/Wales/PHWC-16D7A1/2020|EPI_ISL_577446|2020 ···················· ···················· ···················· ···················· ···················· ····················
L 7117 hCoV-19/Wales/PHWC-16D7B0/2020|EPI_ISL_577447|2020 ···················· ···················· ···················· ···················· ···················· ····················
L 7118 hCoV-19/Wales/PHWC-16D7CF/2020|EPI_ISL_577448|2020 ···················· ···················· ···················· ···················· ···················· ····················
L 7119 hCoV-19/Wales/PHWC-16D7ED/2020|EPI_ISL_577449|2020 ···················· ···················· ···················· ···················· ···················· ····················
L 7120 hCoV-19/Wales/PHWC-16D826/2020|EPI_ISL_577450|2020 ···················· ···················· ···················· ···················· ···················· ····················
L 7121 hCoV-19/Wales/PHWC-16D835/2020|EPI_ISL_577451|2020 ···················· ···················· ···················· ···················· ···················· ····················
L 7122 hCoV-19/Wales/PHWC-16D844/2020|EPI_ISL_577452|2020 ···················· ···················· ···················· ···················· ···················· ····················
L 7123 hCoV-19/Wales/PHWC-16D853/2020|EPI_ISL_577453|2020 ···················· ···················· ···················· ···················· ···················· ····················
L 7124 hCoV-19/Wales/PHWC-16D862/2020|EPI_ISL_577454|2020 ···················· ···················· ···················· ···················· ···················· ····················
L 7125 hCoV-19/Wales/PHWC-16D871/2020|EPI_ISL_577455|2020 ···················· ···················· ···················· ···················· ···················· ····················
L 7126 hCoV-19/Wales/PHWC-16D89F/2020|EPI_ISL_577457|2020 ···················· ···················· ···················· ···················· ···················· ····················
L 7127 hCoV-19/Wales/PHWC-16D96F/2020|EPI_ISL_577470|2020 ···················· ···················· ···················· ···················· ···················· ····················
L 7128 hCoV-19/Wales/PHWC-16D97E/2020|EPI_ISL_577471|2020 ···················· ···················· ···················· ···················· ···················· ····················
L 7129 hCoV-19/Wales/PHWC-16D98D/2020|EPI_ISL_577472|2020 ···················· ···················· ···················· ···················· ···················· ····················
L 7130 hCoV-19/Wales/PHWC-16D99C/2020|EPI_ISL_577473|2020 ···················· ···················· ···················· ···················· ···················· ····················
L 7131 hCoV-19/Wales/PHWC-16D9AB/2020|EPI_ISL_577474|2020 ···················· ···················· ···················· ···················· ···················· ····················
L 7132 hCoV-19/Wales/PHWC-16D9BA/2020|EPI_ISL_577475|2020 ···················· ···················· ···················· ···················· ···················· ····················
L 7133 hCoV-19/Wales/PHWC-16D9D8/2020|EPI_ISL_577476|2020 ···················· ···················· ···················· ···················· ···················· ····················
L 7134 hCoV-19/Wales/PHWC-16D9E7/2020|EPI_ISL_577477|2020 ···················· ···················· ···················· ···················· ···················· ····················
L 7135 hCoV-19/Wales/PHWC-16DA02/2020|EPI_ISL_577478|2020 ···················· ···················· ···················· ···················· ···················· ····················
L 7136 hCoV-19/Wales/PHWC-16DA11/2020|EPI_ISL_577479|2020 ···················· ···················· ···················· ···················· ···················· ····················
L 7137 hCoV-19/USA/LA-EVTL361/2020|EPI_ISL_578767|2020-05 ···················· ···················· ···················· ···················· ···················· ····················
L 7138 hCoV-19/USA/LA-EVTL362/2020|EPI_ISL_578768|2020-05 ···················· ···················· ···················· ···················· ···················· ····················
L 7139 hCoV-19/USA/LA-EVTL363/2020|EPI_ISL_578769|2020-05 ···················· ···················· ···················· ···················· ···················· ····················
L 7140 hCoV-19/USA/LA-EVTL364/2020|EPI_ISL_578770|2020-05 ···················· ···················· ···················· ···················· ···················· ····················
L 7141 hCoV-19/USA/LA-EVTL365/2020|EPI_ISL_578771|2020-05 ···················· ···················· ···················· ···················· ···················· ····················
L 7142 hCoV-19/USA/LA-EVTL366/2020|EPI_ISL_578772|2020-05 ···················· ···················· ···················· ···················· ···················· ····················
L 7143 hCoV-19/USA/LA-EVTL367/2020|EPI_ISL_578773|2020-05 ···················· ···················· ···················· ···················· ···················· ····················
L 7144 hCoV-19/USA/LA-EVTL368/2020|EPI_ISL_578774|2020-06 ···················· ···················· ···················· ···················· ···················· ····················
L 7145 hCoV-19/USA/LA-EVTL369/2020|EPI_ISL_578775|2020-06 ···················· ···················· ···················· ···················· ···················· ····················
L 7146 hCoV-19/USA/LA-EVTL370/2020|EPI_ISL_578776|2020-06 ···················· ···················· ···················· ···················· ···················· ····················
L 7147 hCoV-19/USA/LA-EVTL371/2020|EPI_ISL_578777|2020-06 ···················· ···················· ···················· ···················· ···················· ····················
L 7148 hCoV-19/USA/LA-EVTL372/2020|EPI_ISL_578778|2020-06 ···················· ···················· ···················· ···················· ···················· ····················
L 7149 hCoV-19/USA/LA-EVTL373/2020|EPI_ISL_578779|2020-06 ···················· ···················· ···················· ···················· ···················· ····················
L 7150 hCoV-19/USA/LA-EVTL374/2020|EPI_ISL_578780|2020-06 ···················· ···················· ···················· ···················· ···················· ····················
L 7151 hCoV-19/USA/LA-EVTL375/2020|EPI_ISL_578781|2020-06 ···················· ···················· ···················· ···················· ···················· ····················
L 7152 hCoV-19/USA/LA-EVTL376/2020|EPI_ISL_578782|2020-06 ···················· ···················· ···················· ···················· ···················· ····················
L 7153 hCoV-19/USA/LA-EVTL377/2020|EPI_ISL_578783|2020-06 ···················· ···················· ···················· ···················· ···················· ····················
L 7154 hCoV-19/USA/LA-EVTL378/2020|EPI_ISL_578784|2020-06 ···················· ···················· ···················· ···················· ···················· ····················
L 7155 hCoV-19/USA/LA-EVTL379/2020|EPI_ISL_578785|2020-06 ···················· ···················· ···················· ···················· ···················· ····················
L 7156 hCoV-19/USA/LA-EVTL380/2020|EPI_ISL_578786|2020-06 ···················· ···················· ···················· ···················· ···················· ····················
L 7157 hCoV-19/USA/LA-EVTL381/2020|EPI_ISL_578787|2020-06 ···················· ···················· ···················· ···················· ···················· ····················
L 7158 hCoV-19/USA/LA-EVTL382/2020|EPI_ISL_578788|2020-06 ···················· ···················· ···················· ···················· ···················· ····················
L 7160 hCoV-19/USA/MI-UM-10035752975/2020|EPI_ISL_583301| ···················· ·A·················· ···················· ···················· ···················· ····················
L 7161 hCoV-19/USA/MI-UM-10035733857/2020|EPI_ISL_583302| ···················· ·A·················· ···················· ···················· ···················· ····················
L 7162 hCoV-19/USA/MI-UM-10035757349/2020|EPI_ISL_583303| ············T······· ···················· ···················· ···················· ···················· ····················
L 7163 hCoV-19/USA/MI-UM-10035757569/2020|EPI_ISL_583304| ···················· ···················· ···················· ···················· ···················· ····················
L 7164 hCoV-19/USA/MI-UM-10035711854/2020|EPI_ISL_583305| ···················· ·············A······ ···················· ···················· ···················· ····················
L 7166 hCoV-19/USA/MI-UM-10035747436/2020|EPI_ISL_583307| ···················· ·A····A············· ···················· ···················· ···················· ····················
L 7167 hCoV-19/USA/MI-UM-10035731267/2020|EPI_ISL_583308| ···················· ·A····A············· ···················· ···················· ···················· ····················
L 7168 hCoV-19/USA/MI-UM-10035727656/2020|EPI_ISL_583309| ···················· ·A·················· ···················· ···················· ···················· ····················
L 7169 hCoV-19/Wales/PHWC-16B794/2020|EPI_ISL_573900|2020 ···················· ···················· ···················· ···················· ···················· ····················
L 7170 hCoV-19/Wales/PHWC-16B7A3/2020|EPI_ISL_573901|2020 ···················· ···················· ···················· ···················· ···················· ····················
L 7171 hCoV-19/Wales/PHWC-16B7B2/2020|EPI_ISL_573902|2020 ···················· ···················· ···················· ···················· ···················· ····················
L 7172 hCoV-19/Wales/PHWC-16B7C1/2020|EPI_ISL_573903|2020 ···················· ···················· ···················· ···················· ···················· ····················
L 7173 hCoV-19/Wales/PHWC-16B7EF/2020|EPI_ISL_573904|2020 ···················· ···················· ···················· ···················· ···················· ····················
L 7174 hCoV-19/Wales/PHWC-16B7FE/2020|EPI_ISL_573905|2020 ···················· ···················· ···················· ···················· ···················· ····················
L 7175 hCoV-19/Wales/PHWC-16B80A/2020|EPI_ISL_573906|2020 ···················· ···················· ···················· ···················· ···················· ····················
L 7176 hCoV-19/Wales/PHWC-16B819/2020|EPI_ISL_573907|2020 ···················· ···················· ···················· ···················· ···················· ····················
L 7177 hCoV-19/Wales/PHWC-16B837/2020|EPI_ISL_573908|2020 ···················· ···················· ···················· ···················· ···················· ····················
L 7178 hCoV-19/Wales/PHWC-16B855/2020|EPI_ISL_573909|2020 ···················· ···················· ···················· ···················· ···················· ····················
L 7179 hCoV-19/Indonesia/YO-UGM-10005/2020|EPI_ISL_576128 ···················· ···················· ···················· ···················· ···················· ····················
L 7180 hCoV-19/Indonesia/YO-UGM-10006/2020|EPI_ISL_576130 ···················· ···················· ···················· ···················· ···················· ····················
L 7181 hCoV-19/Australia/QLD1043/2020|EPI_ISL_576131|2020 ···················· ···················· ···················· ···················· ···················· ····················
L 7182 hCoV-19/Australia/QLD1044/2020|EPI_ISL_576132|2020 ···················· ···················· ···················· ···················· ···················· ····················
L 7183 hCoV-19/Australia/QLD1045/2020|EPI_ISL_576133|2020 ···················· ···················· ···················· ···················· ···················· ····················
L 7184 hCoV-19/Australia/QLD1046/2020|EPI_ISL_576134|2020 ···················· ···················· ···················· ···················· ···················· ····················
L 7185 hCoV-19/Australia/QLD1047/2020|EPI_ISL_576135|2020 ···················· ···················· ···················· ···················· ···················· ····················
L 7186 hCoV-19/Australia/QLD1048/2020|EPI_ISL_576136|2020 ···················· ···················· ···················· ···················· ···················· ····················
L 7187 hCoV-19/Australia/QLD1050/2020|EPI_ISL_576137|2020 ···················· ···················· ···················· ···················· ···················· ····················
L 7188 hCoV-19/Australia/QLD1051/2020|EPI_ISL_576138|2020 ···················· ···················· ···················· ···················· ···················· ····················
L 7189 hCoV-19/Australia/QLD1052/2020|EPI_ISL_576139|2020 ···················· ···················· ···················· ···················· ···················· ····················
L 7190 hCoV-19/Ukraine/Lutsk-889/2020|EPI_ISL_576149|2020 ···················· ···················· ···················· ···················· ···················· ····················
L 7191 hCoV-19/USA/DE-DHSS-F918154/2020|EPI_ISL_576150|20 ···················· ···················· ···················· ···················· ···················· ····················
L 7192 hCoV-19/USA/DE-DHSS-F918161/2020|EPI_ISL_576151|20 ···················· ···················· ···················· ···················· ···················· ····················
L 7193 hCoV-19/USA/DE-DHSS-F917991/2020|EPI_ISL_576152|20 ···················· ···················· ···················· ···················· ···················· ····················
L 7194 hCoV-19/USA/DE-DHSS-F917937/2020|EPI_ISL_576153|20 ···················· ···················· ···················· ···················· ···················· ····················
L 7195 hCoV-19/USA/DE-DHSS-F917912/2020|EPI_ISL_576154|20 ···················· ···················· ···················· ···················· ···················· ····················
```

| | | | | | | | | | |
|---|---|---|---|---|---|---|---|---|---|
| L | 7196 | hCoV-19/USA/DE-DHSS-F917890/2020\|EPI_ISL_576155\|20 | .................... | .................... | .................... | .................... | .................... | .................... | |
| L | 7197 | hCoV-19/USA/DE-DHSS-F917932/2020\|EPI_ISL_576156\|20 | .................... | .................... | .................... | .................... | .................... | .................... | |
| L | 7198 | hCoV-19/USA/DE-DHSS-F917849/2020\|EPI_ISL_576157\|20 | .................... | .................... | .................... | .................... | .................... | .................... | |
| L | 7199 | hCoV-19/USA/DE-DHSS-F917986/2020\|EPI_ISL_576158\|20 | .................... | .................... | .................... | .................... | .................... | .................... | |
| L | 7200 | hCoV-19/USA/DE-DHSS-F917888/2020\|EPI_ISL_576160\|20 | .................... | .................... | .................... | .................... | .................... | .................... | |
| L | 7201 | hCoV-19/USA/DE-DHSS-F918105/2020\|EPI_ISL_576161\|20 | .................... | .................... | .................... | .................... | .................... | .................... | |
| L | 7202 | hCoV-19/USA/DE-DHSS-F917966/2020\|EPI_ISL_576162\|20 | .................... | .................... | .................... | .................... | .................... | .................... | |
| L | 7203 | hCoV-19/USA/DE-DHSS-F917896/2020\|EPI_ISL_576164\|20 | .................... | .................... | .................... | .................... | .................... | .................... | |
| L | 7204 | hCoV-19/USA/DE-DHSS-F917892/2020\|EPI_ISL_576165\|20 | .................... | .................... | .................... | .................... | .................... | .................... | |
| L | 7205 | hCoV-19/USA/DE-DHSS-F917974/2020\|EPI_ISL_576166\|20 | .................... | .................... | .................... | .................... | .................... | .................... | |
| L | 7206 | hCoV-19/USA/DE-DHSS-F917893/2020\|EPI_ISL_576167\|20 | .................... | .................... | .................... | .................... | .................... | .................... | |
| L | 7207 | hCoV-19/USA/DE-DHSS-F917785/2020\|EPI_ISL_576169\|20 | .................... | .................... | .................... | .................... | .................... | .................... | |
| L | 7208 | hCoV-19/USA/PA-CDC-8397/2020\|EPI_ISL_576170\|2020-0 | .................... | .................... | .................... | .................... | .................... | .................... | |
| L | 7209 | hCoV-19/USA/PA-CDC-88398/2020\|EPI_ISL_576171\|2020- | .................... | .................... | .................... | .................... | .................... | .................... | |
| L | 7210 | hCoV-19/USA/TN-CDC-8414/2020\|EPI_ISL_576172\|2020-0 | .................... | .................... | .................... | .................... | .................... | .................... | |
| L | 7211 | hCoV-19/USA/FL-CDC-8725/2020\|EPI_ISL_576173\|2020-0 | .................... | .................... | .................... | .................... | .................... | .................... | |
| L | 7212 | hCoV-19/USA/VA-CDC-8752/2020\|EPI_ISL_576174\|2020-0 | .................... | .................... | .................... | .................... | .................... | .................... | |
| L | 7213 | hCoV-19/USA/TN-CDC-8411/2020\|EPI_ISL_576175\|2020-0 | .................... | .................... | .................... | .................... | .................... | .................... | |
| L | 7214 | hCoV-19/USA/TN-CDC-8412/2020\|EPI_ISL_576176\|2020-0 | .................... | .................... | .................... | .................... | .................... | .................... | |
| L | 7215 | hCoV-19/USA/CA-CDC-8726/2020\|EPI_ISL_576177\|2020-0 | .................... | .................... | .................... | .................... | .................... | .................... | |
| L | 7216 | hCoV-19/USA/DC-CDC-8704/2020\|EPI_ISL_576178\|2020-0 | .................... | .................... | .................... | .................... | .................... | .................... | |
| L | 7217 | hCoV-19/USA/WI-CDC-6230/2020\|EPI_ISL_576179\|2020-0 | .................... | .................... | .................... | .................... | .................... | .................... | |
| L | 7218 | hCoV-19/USA/WI-CDC-6231/2020\|EPI_ISL_576180\|2020-0 | .................... | .................... | .................... | .................... | .................... | .................... | |
| L | 7219 | hCoV-19/USA/WI-CDC-6232/2020\|EPI_ISL_576181\|2020-0 | .................... | .................... | .................... | .................... | .................... | .................... | |
| L | 7220 | hCoV-19/USA/WI-CDC-6233/2020\|EPI_ISL_576182\|2020-0 | .................... | .................... | .................... | .................... | .................... | .................... | |
| L | 7221 | hCoV-19/USA/WI-CDC-6234/2020\|EPI_ISL_576183\|2020-0 | .................... | .................... | .................... | .................... | .................... | .................... | |
| L | 7222 | hCoV-19/USA/WI-CDC-6235/2020\|EPI_ISL_576184\|2020-0 | .................... | .................... | .................... | .................... | .................... | .................... | |
| L | 7223 | hCoV-19/USA/WI-CDC-6236/2020\|EPI_ISL_576185\|2020-0 | .................... | .................... | .................... | .................... | .................... | .................... | |
| L | 7224 | hCoV-19/USA/WI-CDC-6237/2020\|EPI_ISL_576186\|2020-0 | .................... | .................... | .................... | .................... | .................... | .................... | |
| L | 7225 | hCoV-19/USA/WI-CDC-6238/2020\|EPI_ISL_576187\|2020-0 | .................... | .................... | .................... | .................... | .................... | .................... | |
| L | 7226 | hCoV-19/USA/WI-CDC-6239/2020\|EPI_ISL_576188\|2020-0 | .................... | .................... | .................... | .................... | .................... | .................... | |
| L | 7227 | hCoV-19/USA/GA-CDC-4447/2020\|EPI_ISL_576189\|2020-0 | .................... | .................... | .................... | .................... | .................... | .................... | |
| L | 7228 | hCoV-19/USA/MI-MDHHS-SC21987/2020\|EPI_ISL_576210\|2 | .................... | .................... | .................... | .................... | .................... | .................... | |
| L | 7229 | hCoV-19/USA/MI-MDHHS-SC21988/2020\|EPI_ISL_576211\|2 | .................... | .................... | .................... | .................... | .................... | .................... | |
| L | 7230 | hCoV-19/USA/MI-MDHHS-SC21989/2020\|EPI_ISL_576212\|2 | .................... | .................... | .................... | .................... | .................... | .................... | |
| L | 7231 | hCoV-19/USA/MI-MDHHS-SC21990/2020\|EPI_ISL_576213\|2 | .................... | .................... | .................... | .................... | .................... | .................... | |
| L | 7232 | hCoV-19/USA/MI-MDHHS-SC21991/2020\|EPI_ISL_576214\|2 | .................... | .................... | .................... | .................... | .................... | .................... | |
| L | 7233 | hCoV-19/USA/MI-MDHHS-SC21992/2020\|EPI_ISL_576215\|2 | .................... | .................... | .................... | .................... | .................... | .................... | |
| L | 7234 | hCoV-19/USA/MI-MDHHS-SC21993/2020\|EPI_ISL_576216\|2 | .................... | .................... | .................... | .................... | .................... | .................... | |
| L | 7235 | hCoV-19/USA/MI-MDHHS-SC21994/2020\|EPI_ISL_576217\|2 | .................... | .................... | .................... | .................... | .................... | .................... | |
| L | 7236 | hCoV-19/USA/MI-MDHHS-SC21995/2020\|EPI_ISL_576218\|2 | .................... | .................... | .................... | .................... | .................... | .................... | |
| L | 7237 | hCoV-19/USA/MI-MDHHS-SC21996/2020\|EPI_ISL_576219\|2 | .................... | .................... | .................... | .................... | .................... | .................... | |
| L | 7238 | hCoV-19/USA/MI-MDHHS-SC21997/2020\|EPI_ISL_576220\|2 | .................... | .................... | .................... | .................... | .................... | .................... | |
| L | 7239 | hCoV-19/USA/MI-MDHHS-SC21998/2020\|EPI_ISL_576221\|2 | .................... | .................... | .................... | .................... | .................... | .................... | |
| L | 7240 | hCoV-19/USA/DE-DHSS-F917945/2020\|EPI_ISL_576223\|20 | .................... | .................... | .................... | .................... | .................... | .................... | |
| L | 7241 | hCoV-19/USA/DE-DHSS-F917930/2020\|EPI_ISL_576224\|20 | .................... | .................... | .................... | .................... | .................... | .................... | |
| L | 7242 | hCoV-19/USA/DE-DHSS-F917934/2020\|EPI_ISL_576225\|20 | .................... | .................... | .................... | .................... | .................... | .................... | |
| L | 7243 | hCoV-19/Wales/PHWC-16D8AE/2020\|EPI_ISL_577458\|2020 | .................... | .................... | .................... | .................... | .................... | .................... | |
| L | 7244 | hCoV-19/Wales/PHWC-16D8BD/2020\|EPI_ISL_577459\|2020 | .................... | .................... | .................... | .................... | .................... | .................... | |
| L | 7245 | hCoV-19/Wales/PHWC-16D8CC/2020\|EPI_ISL_577460\|2020 | .................... | .................... | .................... | .................... | .................... | .................... | |
| L | 7246 | hCoV-19/Wales/PHWC-16D8DB/2020\|EPI_ISL_577461\|2020 | .................... | .................... | .................... | .................... | .................... | .................... | |
| L | 7247 | hCoV-19/Wales/PHWC-16D8EA/2020\|EPI_ISL_577462\|2020 | .................... | .................... | .................... | .................... | .................... | .................... | |
| L | 7248 | hCoV-19/Wales/PHWC-16D8F9/2020\|EPI_ISL_577463\|2020 | .................... | .................... | .................... | .................... | .................... | .................... | |
| L | 7249 | hCoV-19/Wales/PHWC-16D905/2020\|EPI_ISL_577464\|2020 | .................... | .................... | .................... | .................... | .................... | .................... | |
| L | 7250 | hCoV-19/Wales/PHWC-16D914/2020\|EPI_ISL_577465\|2020 | .................... | .................... | .................... | .................... | .................... | .................... | |
| L | 7251 | hCoV-19/Wales/PHWC-16D923/2020\|EPI_ISL_577466\|2020 | .................... | .................... | .................... | .................... | .................... | .................... | |
| L | 7253 | hCoV-19/Wales/PHWC-16D941/2020\|EPI_ISL_577468\|2020 | .................... | .................... | .................... | .................... | .................... | .................... | |
| L | 7255 | hCoV-19/Wales/PHWC-16DA20/2020\|EPI_ISL_577480\|2020 | .................... | .................... | .................... | .................... | .................... | .................... | |
| L | 7256 | hCoV-19/Wales/PHWC-16DA3F/2020\|EPI_ISL_577481\|2020 | .................... | .................... | .................... | .................... | .................... | .................... | |
| L | 7257 | hCoV-19/Wales/PHWC-16DA4E/2020\|EPI_ISL_577482\|2020 | .................... | .................... | .................... | .................... | .................... | .................... | |
| L | 7258 | hCoV-19/Wales/PHWC-16DA7B/2020\|EPI_ISL_577483\|2020 | .................... | .................... | .................... | .................... | .................... | .................... | |
| L | 7259 | hCoV-19/Wales/PHWC-16DA8A/2020\|EPI_ISL_577484\|2020 | .................... | .................... | .................... | .................... | .................... | .................... | |
| L | 7260 | hCoV-19/Wales/PHWC-16DA99/2020\|EPI_ISL_577485\|2020 | .................... | .................... | .................... | .................... | .................... | .................... | |
| L | 7261 | hCoV-19/Wales/PHWC-16DAB7/2020\|EPI_ISL_577486\|2020 | .................... | .................... | .................... | .................... | .................... | .................... | |
| L | 7262 | hCoV-19/Wales/PHWC-16DAC6/2020\|EPI_ISL_577487\|2020 | .................... | .................... | .................... | .................... | .................... | .................... | |
| L | 7263 | hCoV-19/Wales/PHWC-16DAD5/2020\|EPI_ISL_577488\|2020 | .................... | .................... | .................... | .................... | .................... | .................... | |
| L | 7264 | hCoV-19/Wales/PHWC-16DAE4/2020\|EPI_ISL_577489\|2020 | .................... | .................... | .................... | .................... | .................... | .................... | |
| L | 7265 | hCoV-19/Wales/PHWC-16DAF3/2020\|EPI_ISL_577490\|2020 | .................... | .................... | .................... | .................... | .................... | .................... | |
| L | 7266 | hCoV-19/Wales/PHWC-16DB0F/2020\|EPI_ISL_577491\|2020 | .................... | .................... | .................... | .................... | .................... | .................... | |
| L | 7267 | hCoV-19/Wales/PHWC-16DB1E/2020\|EPI_ISL_577492\|2020 | .................... | .................... | .................... | .................... | .................... | .................... | |
| L | 7268 | hCoV-19/Wales/PHWC-16DB2D/2020\|EPI_ISL_577493\|2020 | .................... | .................... | .................... | .................... | .................... | .................... | |
| L | 7271 | hCoV-19/Wales/PHWC-16DB5A/2020\|EPI_ISL_577496\|2020 | .................... | .................... | .................... | .................... | .................... | .................... | |
| L | 7272 | hCoV-19/Wales/PHWC-16DB69/2020\|EPI_ISL_577497\|2020 | .................... | .................... | .................... | .................... | .................... | .................... | |
| L | 7273 | hCoV-19/Wales/PHWC-16DB78/2020\|EPI_ISL_577498\|2020 | .................... | .................... | .................... | .................... | .................... | .................... | |
| L | 7275 | hCoV-19/Wales/PHWC-16DB96/2020\|EPI_ISL_577500\|2020 | .................... | .................... | .................... | .................... | .................... | .................... | |
| L | 7277 | hCoV-19/Wales/PHWC-16DBB4/2020\|EPI_ISL_577502\|2020 | .................... | .................... | .................... | .................... | NNN................. | .................... | 1 |
| L | 7281 | hCoV-19/Wales/PHWC-16DBF0/2020\|EPI_ISL_577506\|2020 | .................... | .................... | .................... | .................... | .................... | .................... | |
| L | 7282 | hCoV-19/Wales/PHWC-16DC0C/2020\|EPI_ISL_577507\|2020 | .................... | .................... | .................... | .................... | .................... | .................... | |
| L | 7285 | hCoV-19/Wales/PHWC-16DC39/2020\|EPI_ISL_577510\|2020 | .................... | .................... | .................... | .................... | .................... | .................... | |
| L | 7289 | hCoV-19/Wales/PHWC-16DCA2/2020\|EPI_ISL_577517\|2020 | .................... | .................... | .................... | .................... | .................... | .................... | |

| | | | | | | | | |
|---|---|---|---|---|---|---|---|---|
| L | 7296 hCoV-19/Wales/PHWC-16DD18/2020\|EPI_ISL_577524\|2020 | ···················· | ···················· | ····················· | ···················· | ···················· | ···················· | |
| L | 7297 hCoV-19/Wales/PHWC-16DD27/2020\|EPI_ISL_577525\|2020 | ···················· | ···················· | ····················· | ···················· | ···················· | ···················· | |
| L | 7300 hCoV-19/Wales/PHWC-16DD54/2020\|EPI_ISL_577528\|2020 | ···················· | ···················· | ····················· | ···················· | ···················· | ···················· | |
| L | 7301 hCoV-19/Wales/PHWC-16DD63/2020\|EPI_ISL_577529\|2020 | ···················· | ···················· | ····················· | ···················· | NNN················· | ···················· | 1 |
| L | 7303 hCoV-19/Wales/PHWC-16DD81/2020\|EPI_ISL_577531\|2020 | ···················· | ···················· | ····················· | ···················· | ···················· | ···················· | |
| L | 7304 hCoV-19/Wales/PHWC-16DD90/2020\|EPI_ISL_577532\|2020 | ···················· | ···················· | ····················· | ···················· | ···················· | ···················· | |
| L | 7305 hCoV-19/Wales/PHWC-16DDAF/2020\|EPI_ISL_577533\|2020 | ···················· | ···················· | ····················· | ···················· | ···················· | ···················· | |
| L | 7307 hCoV-19/Wales/PHWC-16DE06/2020\|EPI_ISL_577539\|2020 | ···················· | ···················· | ····················· | ···················· | ···················· | ···················· | |
| L | 7311 hCoV-19/Wales/PHWC-16DE42/2020\|EPI_ISL_577543\|2020 | ···················· | ···················· | ····················· | ···················· | ···················· | ···················· | |
| L | 7312 hCoV-19/USA/LA-EVTL383/2020\|EPI_ISL_578789\|2020-06 | ···················· | ···················· | ····················· | ···················· | ···················· | ···················· | |
| L | 7313 hCoV-19/USA/LA-EVTL384/2020\|EPI_ISL_578790\|2020-06 | ···················· | ···················· | ····················· | ···················· | ···················· | ···················· | |
| L | 7314 hCoV-19/USA/LA-EVTL385/2020\|EPI_ISL_578791\|2020-06 | ···················· | ···················· | ····················· | ···················· | ···················· | ···················· | |
| L | 7315 hCoV-19/USA/LA-EVTL386/2020\|EPI_ISL_578792\|2020-06 | ···················· | ···················· | ····················· | ···················· | ···················· | ···················· | |
| L | 7316 hCoV-19/USA/LA-EVTL387/2020\|EPI_ISL_578793\|2020-06 | ···················· | ···················· | ····················· | ···················· | ···················· | ···················· | |
| L | 7317 hCoV-19/USA/LA-EVTL388/2020\|EPI_ISL_578794\|2020-06 | ···················· | ···················· | ····················· | ···················· | ···················· | ···················· | |
| L | 7318 hCoV-19/USA/LA-EVTL389/2020\|EPI_ISL_578795\|2020-06 | ···················· | ···················· | ····················· | ···················· | ···················· | ···················· | |
| L | 7319 hCoV-19/USA/LA-EVTL390/2020\|EPI_ISL_578796\|2020-06 | ···················· | ···················· | ····················· | ···················· | ···················· | ···················· | |
| L | 7320 hCoV-19/USA/LA-EVTL391/2020\|EPI_ISL_578797\|2020-06 | ···················· | ···················· | ····················· | ···················· | ···················· | ···················· | |
| L | 7321 hCoV-19/USA/LA-EVTL392/2020\|EPI_ISL_578798\|2020-06 | ···················· | ···················· | ····················· | ···················· | ···················· | ···················· | |
| L | 7322 hCoV-19/USA/LA-EVTL393/2020\|EPI_ISL_578799\|2020-06 | ···················· | ···················· | ····················· | ···················· | ···················· | ···················· | |
| L | 7323 hCoV-19/USA/LA-EVTL394/2020\|EPI_ISL_578800\|2020-06 | ···················· | ···················· | ····················· | ···················· | ···················· | ···················· | |
| L | 7324 hCoV-19/USA/LA-EVTL395/2020\|EPI_ISL_578801\|2020-06 | ···················· | ···················· | ····················· | ···················· | ···················· | ···················· | |
| L | 7325 hCoV-19/USA/LA-EVTL396/2020\|EPI_ISL_578802\|2020-06 | ···················· | ···················· | ····················· | ···················· | ···················· | ···················· | |
| L | 7326 hCoV-19/USA/LA-EVTL397/2020\|EPI_ISL_578803\|2020-06 | ···················· | ···················· | ····················· | ···················· | ···················· | ···················· | |
| L | 7327 hCoV-19/USA/LA-EVTL398/2020\|EPI_ISL_578804\|2020-06 | ···················· | ···················· | ····················· | ···················· | ···················· | ···················· | |
| L | 7328 hCoV-19/USA/LA-EVTL399/2020\|EPI_ISL_578805\|2020-06 | ···················· | ···················· | ····················· | ···················· | ···················· | ···················· | |
| L | 7329 hCoV-19/USA/LA-EVTL400/2020\|EPI_ISL_578806\|2020-06 | ···················· | ···················· | ····················· | ···················· | ···················· | ···················· | |
| L | 7330 hCoV-19/USA/LA-EVTL401/2020\|EPI_ISL_578807\|2020-06 | ···················· | ···················· | ····················· | ···················· | ···················· | ···················· | |
| L | 7331 hCoV-19/USA/LA-EVTL402/2020\|EPI_ISL_578808\|2020-06 | ···················· | ················T··· | ····················· | ···················· | ···················· | ···················· | |
| L | 7332 hCoV-19/USA/LA-EVTL403/2020\|EPI_ISL_578809\|2020-06 | ···················· | ···················· | ····················· | ···················· | ···················· | ···················· | |
| L | 7333 hCoV-19/USA/LA-EVTL404/2020\|EPI_ISL_578810\|2020-06 | ···················· | ···················· | ····················· | ···················· | ···················· | ···················· | |
| L | 7334 hCoV-19/USA/LA-EVTL405/2020\|EPI_ISL_578811\|2020-06 | ···················· | ···················· | ····················· | ···················· | ···················· | ···················· | |
| L | 7335 hCoV-19/USA/LA-EVTL406/2020\|EPI_ISL_578812\|2020-06 | ···················· | ···················· | ····················· | ···················· | ···················· | ···················· | |
| L | 7336 hCoV-19/USA/LA-EVTL407/2020\|EPI_ISL_578813\|2020-06 | ···················· | ···················· | ····················· | ···················· | ···················· | ···················· | |
| L | 7337 hCoV-19/USA/LA-EVTL408/2020\|EPI_ISL_578814\|2020-06 | ···················· | ···················· | ····················· | ···················· | ···················· | ···················· | |
| L | 7338 hCoV-19/USA/LA-EVTL409/2020\|EPI_ISL_578815\|2020-06 | ···················· | ···················· | ····················· | ···················· | ···················· | ···················· | |
| L | 7339 hCoV-19/USA/LA-EVTL418/2020\|EPI_ISL_578824\|2020-07 | ···················· | ·A·················· | ····················· | ···················· | ···················· | ···················· | |
| L | 7340 hCoV-19/USA/LA-EVTL419/2020\|EPI_ISL_578825\|2020-07 | ···················· | ·A·················· | ····················· | ···················· | ···················· | ···················· | |
| L | 7341 hCoV-19/USA/LA-EVTL420/2020\|EPI_ISL_578826\|2020-06 | ···················· | ···················· | ····················· | ···················· | ···················· | ···················· | |
| L | 7342 hCoV-19/USA/LA-EVTL421/2020\|EPI_ISL_578827\|2020-06 | ···················· | ···················· | ····················· | ···················· | ···················· | ···················· | |
| L | 7343 hCoV-19/USA/LA-EVTL422/2020\|EPI_ISL_578828\|2020-06 | ···················· | ···················· | ····················· | ···················· | ···················· | ···················· | |
| L | 7344 hCoV-19/USA/LA-EVTL423/2020\|EPI_ISL_578829\|2020-06 | ···················· | ···················· | ····················· | ···················· | ···················· | ···················· | |
| L | 7345 hCoV-19/USA/LA-EVTL424/2020\|EPI_ISL_578830\|2020-06 | ···················· | ···················· | ····················· | ···················· | ···················· | ···················· | |
| L | 7346 hCoV-19/USA/LA-EVTL425/2020\|EPI_ISL_578831\|2020-07 | ···················· | ·A·················· | ····················· | ···················· | ···················· | ···················· | |
| L | 7347 hCoV-19/USA/LA-EVTL426/2020\|EPI_ISL_578832\|2020-06 | ···················· | ···················· | ····················· | ···················· | ···················· | ···················· | |
| L | 7348 hCoV-19/USA/LA-EVTL427/2020\|EPI_ISL_578833\|2020-06 | ···················· | ···················· | ····················· | ···················· | ···················· | ···················· | |
| L | 7349 hCoV-19/USA/LA-EVTL428/2020\|EPI_ISL_578834\|2020-06 | ···················· | ···················· | ····················· | ···················· | ···················· | ···················· | |
| L | 7350 hCoV-19/USA/LA-EVTL429/2020\|EPI_ISL_578835\|2020-06 | ···················· | ···················· | ····················· | ···················· | ···················· | ···················· | |
| L | 7351 hCoV-19/USA/LA-EVTL430/2020\|EPI_ISL_578836\|2020-06 | ···················· | ···················· | ····················· | ···················· | ···················· | ···················· | |
| L | 7352 hCoV-19/USA/LA-EVTL431/2020\|EPI_ISL_578837\|2020-06 | ···················· | ···················· | ····················· | ···················· | ···················· | ···················· | |
| L | 7353 hCoV-19/USA/LA-EVTL432/2020\|EPI_ISL_578838\|2020-06 | ···················· | ···················· | ····················· | ···················· | ···················· | ···················· | |
| L | 7354 hCoV-19/USA/LA-EVTL433/2020\|EPI_ISL_578839\|2020-06 | ···················· | ···················· | ····················· | ···················· | ···················· | ···················· | |
| L | 7355 hCoV-19/USA/LA-EVTL434/2020\|EPI_ISL_578840\|2020-06 | ···················· | ···················· | ····················· | ···················· | ···················· | ···················· | |
| L | 7356 hCoV-19/USA/LA-EVTL435/2020\|EPI_ISL_578841\|2020-06 | ···················· | ···················· | ····················· | ···················· | ···················· | ···················· | |
| L | 7357 hCoV-19/USA/LA-EVTL436/2020\|EPI_ISL_578842\|2020-06 | ···················· | ···················· | ····················· | ···················· | ···················· | ···················· | |
| L | 7358 hCoV-19/USA/LA-EVTL439/2020\|EPI_ISL_578843\|2020-06 | ···················· | ···················· | ····················· | ···················· | ···················· | ···················· | |
| L | 7359 hCoV-19/USA/LA-EVTL440/2020\|EPI_ISL_578844\|2020-06 | ···················· | ···················· | ····················· | ···················· | ···················· | ···················· | |
| L | 7360 hCoV-19/USA/LA-EVTL441/2020\|EPI_ISL_578845\|2020-06 | ···················· | ···················· | ····················· | ···················· | ···················· | ···················· | |
| L | 7361 hCoV-19/USA/LA-EVTL442/2020\|EPI_ISL_578846\|2020-06 | ···················· | ···················· | ····················· | ···················· | ···················· | ···················· | |
| L | 7362 hCoV-19/USA/LA-EVTL443/2020\|EPI_ISL_578847\|2020-07 | ···················· | ···················· | ····················· | ···················· | ···················· | ···················· | |
| L | 7363 hCoV-19/USA/LA-EVTL444/2020\|EPI_ISL_578848\|2020-06 | ···················· | ···················· | ····················· | ···················· | ···················· | ···················· | |
| L | 7364 hCoV-19/USA/LA-EVTL445/2020\|EPI_ISL_578849\|2020-07 | ···················· | ···················· | ····················· | ···················· | ···················· | ···················· | |
| L | 7365 hCoV-19/USA/LA-EVTL446/2020\|EPI_ISL_578850\|2020-07 | ···················· | ···················· | ····················· | ···················· | ···················· | ···················· | |
| L | 7366 hCoV-19/USA/LA-EVTL447/2020\|EPI_ISL_578851\|2020-06 | ···················· | ···················· | ····················· | ···················· | ···················· | ···················· | |
| L | 7367 hCoV-19/USA/LA-EVTL448/2020\|EPI_ISL_578852\|2020-06 | ···················· | ···················· | ····················· | ···················· | ···················· | ···················· | |
| L | 7368 hCoV-19/USA/LA-EVTL449/2020\|EPI_ISL_578853\|2020-06 | ···················· | ···················· | ····················· | ···················· | ···················· | ···················· | |
| L | 7369 hCoV-19/USA/LA-EVTL450/2020\|EPI_ISL_578854\|2020-07 | ···················· | ···················· | ····················· | ···················· | ···················· | ···················· | |
| L | 7370 hCoV-19/USA/LA-EVTL451/2020\|EPI_ISL_578855\|2020-06 | ···················· | ···················· | ····················· | ···················· | ···················· | ···················· | |
| L | 7371 hCoV-19/USA/LA-EVTL452/2020\|EPI_ISL_578856\|2020-06 | ···················· | ···················· | ····················· | ···················· | ···················· | ···················· | |
| L | 7372 hCoV-19/USA/LA-EVTL453/2020\|EPI_ISL_578857\|2020-06 | ···················· | ···················· | ····················· | ···················· | ···················· | ···················· | |
| L | 7373 hCoV-19/USA/LA-EVTL454/2020\|EPI_ISL_578858\|2020-06 | ···················· | ·············A······ | ····················· | ···················· | ···················· | ···················· | |
| L | 7374 hCoV-19/USA/LA-EVTL455/2020\|EPI_ISL_578859\|2020-06 | ···················· | ···················· | ····················· | ···················· | ···················· | ···················· | |
| L | 7375 hCoV-19/USA/LA-EVTL456/2020\|EPI_ISL_578860\|2020-06 | ···················· | ···················· | ····················· | ···················· | ···················· | ···················· | |
| L | 7376 hCoV-19/USA/LA-EVTL457/2020\|EPI_ISL_578861\|2020-07 | ···················· | ···················· | ····················· | ···················· | ···················· | ···················· | |
| L | 7377 hCoV-19/USA/LA-EVTL458/2020\|EPI_ISL_578862\|2020-06 | ···················· | ···················· | ····················· | ···················· | ···················· | ···················· | |
| L | 7378 hCoV-19/USA/LA-EVTL459/2020\|EPI_ISL_578863\|2020-06 | ···················· | ···················· | ···T················· | ···················· | ···················· | ···················· | |
| L | 7379 hCoV-19/USA/LA-EVTL460/2020\|EPI_ISL_578864\|2020-06 | ···················· | ···················· | ····················· | ···················· | ···················· | ···················· | |
| L | 7380 hCoV-19/USA/LA-EVTL461/2020\|EPI_ISL_578865\|2020-06 | ···················· | ···················· | ····················· | ···················· | ···················· | ···················· | |
| L | 7381 hCoV-19/USA/LA-EVTL466/2020\|EPI_ISL_578870\|2020-06 | ···················· | ···················· | ····················· | ···················· | ···················· | ···················· | |
| L | 7382 hCoV-19/USA/LA-EVTL467/2020\|EPI_ISL_578871\|2020-06 | ···················· | ···················· | ····················· | ···················· | ···················· | ···················· | |

| | | | | | | | | |
|---|---|---|---|---|---|---|---|---|
| L | 7383 | hCoV-19/USA/LA-EVTL468/2020\|EPI_ISL_578872\|2020-06 | ................... | ................... | .................... | ................... | ................... | ................... |
| L | 7384 | hCoV-19/USA/LA-EVTL469/2020\|EPI_ISL_578873\|2020-06 | ................... | ................... | .................... | ................... | ................... | ................... |
| L | 7385 | hCoV-19/USA/LA-EVTL470/2020\|EPI_ISL_578874\|2020-06 | ................... | ................... | .................... | ................... | ................... | ................... |
| L | 7386 | hCoV-19/USA/LA-EVTL471/2020\|EPI_ISL_578875\|2020-06 | ................... | ................... | .................... | ................... | ................... | ................... |
| L | 7387 | hCoV-19/USA/LA-EVTL472/2020\|EPI_ISL_578876\|2020-06 | ................... | ................... | .................... | ................... | ................... | ................... |
| L | 7388 | hCoV-19/USA/LA-EVTL473/2020\|EPI_ISL_578877\|2020-06 | ................... | ................... | .................... | ................... | ................... | ................... |
| L | 7389 | hCoV-19/USA/LA-EVTL474/2020\|EPI_ISL_578878\|2020-06 | ................... | ................... | .................... | ................... | ................... | ................... |
| L | 7390 | hCoV-19/USA/LA-EVTL475/2020\|EPI_ISL_578879\|2020-06 | ................... | ................... | .................... | ................... | ................... | ................... |
| L | 7391 | hCoV-19/USA/LA-EVTL476/2020\|EPI_ISL_578880\|2020-06 | ................... | ................... | .................... | ................... | ................... | ................... |
| L | 7392 | hCoV-19/USA/LA-EVTL477/2020\|EPI_ISL_578881\|2020-06 | ................... | ................... | .................... | ................... | ................... | ................... |
| L | 7393 | hCoV-19/USA/LA-EVTL478/2020\|EPI_ISL_578882\|2020-06 | ................... | ................... | .................... | ................... | ................... | ................... |
| L | 7394 | hCoV-19/USA/LA-EVTL479/2020\|EPI_ISL_578883\|2020-06 | ................... | ................... | .................... | ................... | ................... | ................... |
| L | 7395 | hCoV-19/USA/LA-EVTL480/2020\|EPI_ISL_578884\|2020-06 | ................... | ................... | .................... | ................... | ................... | ................... |
| L | 7396 | hCoV-19/USA/LA-EVTL481/2020\|EPI_ISL_578885\|2020-06 | ................... | ................... | .................... | ................... | ................... | ................... |
| L | 7397 | hCoV-19/USA/LA-EVTL482/2020\|EPI_ISL_578886\|2020-06 | ................... | ................... | .................... | ................... | ................... | ................... |
| L | 7398 | hCoV-19/USA/LA-EVTL483/2020\|EPI_ISL_578887\|2020-06 | ................... | ................... | .................... | ................... | ................... | ................... |
| L | 7399 | hCoV-19/Spain/PV-IBV-98007632/2020\|EPI_ISL_582067\| | ................... | ................... | .................... | ................... | ................... | ................... |
| L | 7400 | hCoV-19/Spain/PV-IBV-98007636/2020\|EPI_ISL_582068\| | ................... | ................... | .................... | ................... | ................... | ................... |
| L | 7401 | hCoV-19/Spain/PV-IBV-98007644/2020\|EPI_ISL_582069\| | ................... | ................... | .................... | ................... | ................... | ................... |
| L | 7402 | hCoV-19/Spain/PV-IBV-98007645/2020\|EPI_ISL_582070\| | ................... | ................... | .................... | ................... | ................... | ................... |
| L | 7403 | hCoV-19/Spain/PV-IBV-98007646/2020\|EPI_ISL_582071\| | ................... | ................... | .................... | ................... | ................... | ................... |
| L | 7404 | hCoV-19/Spain/PV-IBV-98007655/2020\|EPI_ISL_582072\| | ................... | ................... | .................... | ................... | ................... | ................... |
| L | 7405 | hCoV-19/Spain/PV-IBV-98007664/2020\|EPI_ISL_582073\| | ................... | ................... | .................... | ................... | ................... | ................... |
| L | 7406 | hCoV-19/Spain/PV-IBV-98007668/2020\|EPI_ISL_582074\| | ................... | ................... | .................... | ................... | ................... | ................... |
| L | 7407 | hCoV-19/Spain/PV-IBV-98007669/2020\|EPI_ISL_582075\| | ................... | ................... | .................... | ................... | ................... | ................... |
| L | 7408 | hCoV-19/Spain/PV-IBV-98007670/2020\|EPI_ISL_582076\| | ................... | ................... | .................... | ................... | ................... | ................... |
| L | 7409 | hCoV-19/Spain/PV-IBV-98007677/2020\|EPI_ISL_582077\| | ................... | ................... | .................... | ................... | ................... | ................... |
| L | 7410 | hCoV-19/Spain/PV-IBV-98007679/2020\|EPI_ISL_582078\| | ................... | ................... | .................... | ................... | ................... | ................... |
| L | 7411 | hCoV-19/Spain/PV-IBV-98007684/2020\|EPI_ISL_582079\| | ................... | ................... | .................... | ................... | ................... | ................... |
| L | 7412 | hCoV-19/Spain/PV-IBV-98007689/2020\|EPI_ISL_582080\| | ................... | ................... | .................... | ................... | ................... | ................... |
| L | 7413 | hCoV-19/Spain/PV-IBV-98007694/2020\|EPI_ISL_582081\| | ................... | ................... | .................... | ................... | ................... | ................... |
| L | 7414 | hCoV-19/Spain/PV-IBV-98007696/2020\|EPI_ISL_582082\| | ................... | ................... | .................... | ................... | ................... | ................... |
| L | 7415 | hCoV-19/Spain/PV-IBV-98007702/2020\|EPI_ISL_582083\| | ................... | ................... | .................... | ................... | ................... | ................... |
| L | 7416 | hCoV-19/Spain/PV-IBV-98007704/2020\|EPI_ISL_582084\| | ................... | ................... | .................... | ................... | ................... | ................... |
| L | 7418 | hCoV-19/Spain/PV-IBV-98007709/2020\|EPI_ISL_582086\| | ................... | ................... | .................... | ................... | ................... | ................... |
| L | 7419 | hCoV-19/Spain/PV-IBV-98007710/2020\|EPI_ISL_582087\| | ................... | ................... | .................... | ................... | ................... | ................... |
| L | 7420 | hCoV-19/Spain/PV-IBV-98007716/2020\|EPI_ISL_582088\| | ................... | ................... | .................... | ................... | ................... | ................... |
| L | 7421 | hCoV-19/Spain/PV-IBV-98007717/2020\|EPI_ISL_582089\| | ................... | ................... | .................... | ................... | ................... | ................... |
| L | 7422 | hCoV-19/Spain/PV-IBV-98007725/2020\|EPI_ISL_582090\| | ................... | ................... | .................... | ................... | ................... | ................... |
| L | 7423 | hCoV-19/USA/MI-UM-10035923574/2020\|EPI_ISL_583398\| | ................... | ................... | .................... | ................... | ................... | ................... |
| L | 7425 | hCoV-19/USA/MI-QDX-593/2020\|EPI_ISL_571331\|2020-03 | ................... | ................... | .................... | ................... | ................... | ................... |
| L | 7426 | hCoV-19/USA/MI-QDX-595/2020\|EPI_ISL_571332\|2020-03 | ................... | ................... | .................... | ................... | ................... | ................... |
| L | 7427 | hCoV-19/USA/MI-QDX-596/2020\|EPI_ISL_571333\|2020-03 | ................... | ................... | .................... | ................... | ................... | ................... |
| L | 7428 | hCoV-19/USA/MI-QDX-598/2020\|EPI_ISL_571334\|2020-03 | ................... | ................... | .................... | ................... | ................... | ................... |
| L | 7429 | hCoV-19/USA/MI-QDX-600/2020\|EPI_ISL_571335\|2020-03 | ................... | ................... | .................... | ................... | ................... | ................... |
| L | 7430 | hCoV-19/USA/MI-QDX-601/2020\|EPI_ISL_571336\|2020-03 | ................... | ................... | .................... | ................... | ................... | ................... |
| L | 7431 | hCoV-19/USA/MI-QDX-602/2020\|EPI_ISL_571337\|2020-03 | ................... | ................... | .................... | ................... | ................... | ................... |
| L | 7432 | hCoV-19/USA/MI-QDX-603/2020\|EPI_ISL_571338\|2020-03 | ................... | ................... | .................... | ................... | ................... | ................... |
| L | 7433 | hCoV-19/USA/MI-QDX-608/2020\|EPI_ISL_571339\|2020-03 | ................... | ................... | .................... | ................... | ................... | ................... |
| L | 7434 | hCoV-19/USA/MI-QDX-611/2020\|EPI_ISL_571340\|2020-03 | ................... | ................... | .................... | ................... | ................... | ................... |
| L | 7435 | hCoV-19/USA/MI-QDX-612/2020\|EPI_ISL_571341\|2020-03 | ................... | ................... | .................... | ................... | ................... | ................... |
| L | 7436 | hCoV-19/USA/MI-QDX-613/2020\|EPI_ISL_571342\|2020-03 | ................... | ................... | .................... | ................... | ................... | ................... |
| L | 7437 | hCoV-19/USA/MI-QDX-614/2020\|EPI_ISL_571343\|2020-03 | ................... | ................... | .................... | ................... | ................... | ................... |
| L | 7438 | hCoV-19/USA/MI-QDX-615/2020\|EPI_ISL_571344\|2020-03 | ................... | ................... | .................... | ................... | ................... | ................... |
| L | 7439 | hCoV-19/USA/MI-QDX-616/2020\|EPI_ISL_571345\|2020-03 | ................... | ................... | .................... | ................... | ................... | ................... |
| L | 7440 | hCoV-19/USA/MI-QDX-619/2020\|EPI_ISL_571346\|2020-03 | ................... | ................... | ...........T........ | ................... | ................... | ................... |
| L | 7441 | hCoV-19/USA/MI-QDX-620/2020\|EPI_ISL_571347\|2020-03 | ................... | ................... | .................... | ................... | ................... | ................... |
| L | 7442 | hCoV-19/USA/MI-QDX-621/2020\|EPI_ISL_571348\|2020-03 | ................... | ................... | .................... | ................... | ................... | ................... |
| L | 7443 | hCoV-19/USA/MI-QDX-622/2020\|EPI_ISL_571349\|2020-03 | ................... | ................... | .................... | ................... | ................... | ................... |
| L | 7444 | hCoV-19/USA/MI-QDX-623/2020\|EPI_ISL_571350\|2020-03 | ................... | ................... | .................... | ................... | ................... | ................... |
| L | 7445 | hCoV-19/USA/MI-QDX-624/2020\|EPI_ISL_571351\|2020-03 | ................... | ................... | .................... | ................... | ................... | ................... |
| L | 7446 | hCoV-19/USA/MI-QDX-625/2020\|EPI_ISL_571352\|2020-03 | ................... | ................... | .................... | ................... | ................... | ................... |
| L | 7447 | hCoV-19/USA/MI-QDX-626/2020\|EPI_ISL_571353\|2020-03 | ................... | ................... | .................... | ................... | ................... | ................... |
| L | 7448 | hCoV-19/USA/MI-QDX-627/2020\|EPI_ISL_571354\|2020-03 | ................... | ................... | .................... | ................... | ................... | ................... |
| L | 7449 | hCoV-19/USA/MI-QDX-628/2020\|EPI_ISL_571355\|2020-03 | ................... | ................... | .................... | ................... | ................... | ................... |
| L | 7450 | hCoV-19/USA/MI-QDX-629/2020\|EPI_ISL_571356\|2020-03 | ................... | ................... | .................... | ................... | ................... | ................... |
| L | 7451 | hCoV-19/USA/MI-QDX-630/2020\|EPI_ISL_571357\|2020-03 | ................... | ................... | .................... | ................... | ................... | ................... |
| L | 7452 | hCoV-19/USA/MI-QDX-632/2020\|EPI_ISL_571358\|2020-03 | ................... | ................... | .................... | ................... | ................... | ................... |
| L | 7453 | hCoV-19/USA/MI-QDX-634/2020\|EPI_ISL_571359\|2020-03 | ................... | ................... | .................... | ................... | ................... | ................... |
| L | 7454 | hCoV-19/USA/MI-QDX-635/2020\|EPI_ISL_571360\|2020-03 | ................... | ................... | .................... | ................... | ................... | ................... |
| L | 7455 | hCoV-19/USA/MI-QDX-636/2020\|EPI_ISL_571361\|2020-03 | ................... | ................... | .................... | ................... | ................... | ................... |
| L | 7456 | hCoV-19/USA/MI-QDX-637/2020\|EPI_ISL_571362\|2020-03 | ................... | ................... | .................... | ................... | ................... | ................... |
| L | 7457 | hCoV-19/USA/MI-QDX-638/2020\|EPI_ISL_571363\|2020-03 | ................... | ................... | .................... | ................... | ................... | ................... |
| L | 7458 | hCoV-19/USA/MI-QDX-639/2020\|EPI_ISL_571364\|2020-03 | ................... | ................... | .................... | ................... | ................... | ................... |
| L | 7459 | hCoV-19/USA/MI-QDX-640/2020\|EPI_ISL_571365\|2020-03 | ................... | ................... | .................... | ................... | ................... | ................... |
| L | 7460 | hCoV-19/USA/MI-QDX-641/2020\|EPI_ISL_571366\|2020-03 | ................... | ................... | .................... | ................... | ................... | ................... |
| L | 7461 | hCoV-19/USA/MI-QDX-642/2020\|EPI_ISL_571367\|2020-03 | ................... | ................... | .................... | ................... | ................... | ................... |
| L | 7462 | hCoV-19/USA/MI-QDX-643/2020\|EPI_ISL_571368\|2020-03 | ................... | ................... | .................... | ................... | ................... | ................... |
| L | 7463 | hCoV-19/USA/MI-QDX-644/2020\|EPI_ISL_571369\|2020-03 | ................... | ................... | .................... | ................... | ................... | ................... |
| L | 7464 | hCoV-19/USA/MI-QDX-645/2020\|EPI_ISL_571370\|2020-03 | ................... | ................... | .................... | ................... | ................... | ................... |

<table>
<tr><td>L</td><td>7465</td><td>hCoV-19/USA/MI-QDX-646/2020|EPI_ISL_571371|2020-03</td><td>····················</td><td>····················</td><td>·····················</td><td>····················</td><td>····················</td><td>····················</td></tr>
<tr><td>L</td><td>7466</td><td>hCoV-19/USA/MI-QDX-647/2020|EPI_ISL_571372|2020-03</td><td>····················</td><td>····················</td><td>·····················</td><td>····················</td><td>····················</td><td>····················</td></tr>
<tr><td>L</td><td>7467</td><td>hCoV-19/USA/MI-QDX-648/2020|EPI_ISL_571373|2020-03</td><td>····················</td><td>····················</td><td>·····················</td><td>····················</td><td>····················</td><td>····················</td></tr>
<tr><td>L</td><td>7468</td><td>hCoV-19/USA/MI-QDX-651/2020|EPI_ISL_571374|2020-03</td><td>····················</td><td>····················</td><td>·····················</td><td>····················</td><td>····················</td><td>····················</td></tr>
<tr><td>L</td><td>7469</td><td>hCoV-19/USA/MI-QDX-652/2020|EPI_ISL_571375|2020-03</td><td>····················</td><td>····················</td><td>·····················</td><td>····················</td><td>····················</td><td>····················</td></tr>
<tr><td>L</td><td>7470</td><td>hCoV-19/USA/MI-QDX-653/2020|EPI_ISL_571376|2020-03</td><td>····················</td><td>····················</td><td>·····················</td><td>····················</td><td>····················</td><td>····················</td></tr>
<tr><td>L</td><td>7471</td><td>hCoV-19/USA/MI-QDX-654/2020|EPI_ISL_571377|2020-03</td><td>····················</td><td>····················</td><td>·····················</td><td>····················</td><td>····················</td><td>····················</td></tr>
<tr><td>L</td><td>7472</td><td>hCoV-19/USA/MI-QDX-655/2020|EPI_ISL_571378|2020-03</td><td>····················</td><td>····················</td><td>·····················</td><td>····················</td><td>····················</td><td>····················</td></tr>
<tr><td>L</td><td>7473</td><td>hCoV-19/USA/MI-QDX-656/2020|EPI_ISL_571379|2020-03</td><td>····················</td><td>····················</td><td>·····················</td><td>····················</td><td>····················</td><td>····················</td></tr>
<tr><td>L</td><td>7474</td><td>hCoV-19/USA/MI-QDX-657/2020|EPI_ISL_571380|2020-03</td><td>····················</td><td>····················</td><td>·····················</td><td>····················</td><td>····················</td><td>····················</td></tr>
<tr><td>L</td><td>7475</td><td>hCoV-19/USA/MO-QDX-524/2020|EPI_ISL_571381|2020-03</td><td>····················</td><td>····················</td><td>·····················</td><td>····················</td><td>····················</td><td>····················</td></tr>
<tr><td>L</td><td>7476</td><td>hCoV-19/USA/MO-QDX-530/2020|EPI_ISL_571382|2020-03</td><td>····················</td><td>····················</td><td>·····················</td><td>····················</td><td>····················</td><td>····················</td></tr>
<tr><td>L</td><td>7477</td><td>hCoV-19/USA/MO-QDX-531/2020|EPI_ISL_571383|2020-03</td><td>····················</td><td>····················</td><td>·····················</td><td>····················</td><td>····················</td><td>····················</td></tr>
<tr><td>L</td><td>7478</td><td>hCoV-19/USA/MO-QDX-533/2020|EPI_ISL_571384|2020-03</td><td>····················</td><td>····················</td><td>·····················</td><td>····················</td><td>····················</td><td>····················</td></tr>
<tr><td>L</td><td>7479</td><td>hCoV-19/USA/MO-QDX-534/2020|EPI_ISL_571385|2020-03</td><td>····················</td><td>····················</td><td>·····················</td><td>····················</td><td>····················</td><td>····················</td></tr>
<tr><td>L</td><td>7480</td><td>hCoV-19/USA/MO-QDX-535/2020|EPI_ISL_571386|2020-03</td><td>····················</td><td>····················</td><td>·····················</td><td>····················</td><td>····················</td><td>····················</td></tr>
<tr><td>L</td><td>7481</td><td>hCoV-19/USA/MO-QDX-536/2020|EPI_ISL_571387|2020-03</td><td>····················</td><td>····················</td><td>·····················</td><td>····················</td><td>····················</td><td>····················</td></tr>
<tr><td>L</td><td>7482</td><td>hCoV-19/USA/NH-QDX-618/2020|EPI_ISL_571389|2020-03</td><td>····················</td><td>····················</td><td>·····················</td><td>····················</td><td>····················</td><td>····················</td></tr>
<tr><td>L</td><td>7483</td><td>hCoV-19/USA/NV-QDX-538/2020|EPI_ISL_571390|2020-03</td><td>····················</td><td>····················</td><td>·····················</td><td>····················</td><td>····················</td><td>····················</td></tr>
<tr><td>L</td><td>7484</td><td>hCoV-19/USA/NV-QDX-539/2020|EPI_ISL_571391|2020-03</td><td>····················</td><td>····················</td><td>·····················</td><td>····················</td><td>····················</td><td>····················</td></tr>
<tr><td>L</td><td>7485</td><td>hCoV-19/USA/NV-QDX-540/2020|EPI_ISL_571392|2020-03</td><td>····················</td><td>····················</td><td>·····················</td><td>····················</td><td>····················</td><td>····················</td></tr>
<tr><td>L</td><td>7486</td><td>hCoV-19/USA/NV-QDX-541/2020|EPI_ISL_571393|2020-03</td><td>····················</td><td>····················</td><td>·····················</td><td>····················</td><td>····················</td><td>····················</td></tr>
<tr><td>L</td><td>7487</td><td>hCoV-19/USA/NV-QDX-542/2020|EPI_ISL_571394|2020-03</td><td>····················</td><td>····················</td><td>·····················</td><td>····················</td><td>····················</td><td>····················</td></tr>
<tr><td>L</td><td>7488</td><td>hCoV-19/USA/NV-QDX-543/2020|EPI_ISL_571395|2020-03</td><td>····················</td><td>····················</td><td>·····················</td><td>····················</td><td>····················</td><td>····················</td></tr>
<tr><td>L</td><td>7489</td><td>hCoV-19/USA/NV-QDX-544/2020|EPI_ISL_571396|2020-03</td><td>····················</td><td>····················</td><td>·····················</td><td>····················</td><td>····················</td><td>····················</td></tr>
<tr><td>L</td><td>7490</td><td>hCoV-19/USA/WA-QDX-522/2020|EPI_ISL_571398|2020-03</td><td>····················</td><td>····················</td><td>·····················</td><td>····················</td><td>····················</td><td>····················</td></tr>
<tr><td>L</td><td>7491</td><td>hCoV-19/USA/WA-QDX-526/2020|EPI_ISL_571399|2020-03</td><td>····················</td><td>····················</td><td>·····················</td><td>····················</td><td>····················</td><td>····················</td></tr>
<tr><td>L</td><td>7492</td><td>hCoV-19/Scotland/QEUH-9AEAC1/2020|EPI_ISL_573800|2</td><td>····················</td><td>····················</td><td>·····················</td><td>····················</td><td>····················</td><td>····················</td></tr>
<tr><td>L</td><td>7493</td><td>hCoV-19/Scotland/QEUH-9AEAD0/2020|EPI_ISL_573801|2</td><td>····················</td><td>····················</td><td>·····················</td><td>····················</td><td>····················</td><td>····················</td></tr>
<tr><td>L</td><td>7494</td><td>hCoV-19/Scotland/QEUH-9AEAEF/2020|EPI_ISL_573802|2</td><td>····················</td><td>····················</td><td>·····················</td><td>····················</td><td>····················</td><td>····················</td></tr>
<tr><td>L</td><td>7495</td><td>hCoV-19/Scotland/QEUH-9AEAFE/2020|EPI_ISL_573803|2</td><td>····················</td><td>····················</td><td>·····················</td><td>····················</td><td>····················</td><td>····················</td></tr>
<tr><td>L</td><td>7496</td><td>hCoV-19/Scotland/QEUH-9AEC34/2020|EPI_ISL_573804|2</td><td>····················</td><td>····················</td><td>·····················</td><td>····················</td><td>····················</td><td>····················</td></tr>
<tr><td>L</td><td>7497</td><td>hCoV-19/Scotland/QEUH-9AF064/2020|EPI_ISL_573805|2</td><td>····················</td><td>····················</td><td>·····················</td><td>····················</td><td>····················</td><td>····················</td></tr>
<tr><td>L</td><td>7498</td><td>hCoV-19/Scotland/QEUH-9B3CA5/2020|EPI_ISL_573806|2</td><td>····················</td><td>····················</td><td>·····················</td><td>····················</td><td>····················</td><td>····················</td></tr>
<tr><td>L</td><td>7499</td><td>hCoV-19/Scotland/QEUH-9B3E27/2020|EPI_ISL_573807|2</td><td>····················</td><td>····················</td><td>·····················</td><td>····················</td><td>····················</td><td>····················</td></tr>
<tr><td>L</td><td>7500</td><td>hCoV-19/Scotland/QEUH-9B3E54/2020|EPI_ISL_573808|2</td><td>····················</td><td>·A··················</td><td>·····················</td><td>····················</td><td>····················</td><td>··A·················</td></tr>
<tr><td>L</td><td>7501</td><td>hCoV-19/Scotland/QEUH-9B3F7F/2020|EPI_ISL_573809|2</td><td>····················</td><td>·A··················</td><td>·····················</td><td>····················</td><td>····················</td><td>··A·················</td></tr>
<tr><td>L</td><td>7502</td><td>hCoV-19/Scotland/QEUH-9B44CA/2020|EPI_ISL_573810|2</td><td>····················</td><td>····················</td><td>·····················</td><td>····················</td><td>····················</td><td>····················</td></tr>
<tr><td>L</td><td>7503</td><td>hCoV-19/Scotland/QEUH-9BA181/2020|EPI_ISL_573811|2</td><td>····················</td><td>····················</td><td>·····················</td><td>····················</td><td>····················</td><td>····················</td></tr>
<tr><td>L</td><td>7504</td><td>hCoV-19/Scotland/QEUH-9BA400/2020|EPI_ISL_573812|2</td><td>····················</td><td>····················</td><td>·····················</td><td>····················</td><td>····················</td><td>····················</td></tr>
<tr><td>L</td><td>7505</td><td>hCoV-19/Scotland/QEUH-9BA44C/2020|EPI_ISL_573813|2</td><td>····················</td><td>····················</td><td>·····················</td><td>····················</td><td>····················</td><td>····················</td></tr>
<tr><td>L</td><td>7506</td><td>hCoV-19/Scotland/QEUH-9BA45B/2020|EPI_ISL_573814|2</td><td>····················</td><td>····················</td><td>·····················</td><td>····················</td><td>····················</td><td>····················</td></tr>
<tr><td>L</td><td>7507</td><td>hCoV-19/Scotland/QEUH-9BA50D/2020|EPI_ISL_573815|2</td><td>····················</td><td>····················</td><td>·····················</td><td>····················</td><td>····················</td><td>····················</td></tr>
<tr><td>L</td><td>7508</td><td>hCoV-19/Scotland/QEUH-9BA585/2020|EPI_ISL_573816|2</td><td>····················</td><td>····················</td><td>·····················</td><td>····················</td><td>····················</td><td>····················</td></tr>
<tr><td>L</td><td>7509</td><td>hCoV-19/Scotland/QEUH-9BA5D0/2020|EPI_ISL_573817|2</td><td>····················</td><td>····················</td><td>·····················</td><td>····················</td><td>····················</td><td>····················</td></tr>
<tr><td>L</td><td>7510</td><td>hCoV-19/Scotland/QEUH-9BA5EF/2020|EPI_ISL_573818|2</td><td>····················</td><td>····················</td><td>·····················</td><td>····················</td><td>····················</td><td>····················</td></tr>
<tr><td>L</td><td>7511</td><td>hCoV-19/Scotland/QEUH-9BA60A/2020|EPI_ISL_573819|2</td><td>····················</td><td>····················</td><td>·····················</td><td>····················</td><td>····················</td><td>····················</td></tr>
<tr><td>L</td><td>7512</td><td>hCoV-19/Scotland/QEUH-9BA628/2020|EPI_ISL_573820|2</td><td>····················</td><td>····················</td><td>·····················</td><td>····················</td><td>····················</td><td>····················</td></tr>
<tr><td>L</td><td>7513</td><td>hCoV-19/Scotland/QEUH-9BA655/2020|EPI_ISL_573821|2</td><td>····················</td><td>····················</td><td>·····················</td><td>····················</td><td>····················</td><td>····················</td></tr>
<tr><td>L</td><td>7514</td><td>hCoV-19/Scotland/QEUH-9BA664/2020|EPI_ISL_573822|2</td><td>····················</td><td>····················</td><td>·····················</td><td>····················</td><td>····················</td><td>····················</td></tr>
<tr><td>L</td><td>7515</td><td>hCoV-19/Scotland/QEUH-9BA682/2020|EPI_ISL_573823|2</td><td>····················</td><td>·····C··············</td><td>·····················</td><td>····················</td><td>····················</td><td>····················</td></tr>
<tr><td>L</td><td>7516</td><td>hCoV-19/Scotland/QEUH-9BA6CE/2020|EPI_ISL_573824|2</td><td>····················</td><td>····················</td><td>·····················</td><td>····················</td><td>····T···············</td><td>····················</td></tr>
<tr><td>L</td><td>7517</td><td>hCoV-19/Scotland/QEUH-9BA6DD/2020|EPI_ISL_573825|2</td><td>····················</td><td>·A··················</td><td>·····················</td><td>····················</td><td>····················</td><td>··A·················</td></tr>
<tr><td>L</td><td>7518</td><td>hCoV-19/Scotland/QEUH-9BA6FB/2020|EPI_ISL_573826|2</td><td>····················</td><td>·····C··············</td><td>·····················</td><td>····················</td><td>····················</td><td>····················</td></tr>
<tr><td>L</td><td>7519</td><td>hCoV-19/Scotland/QEUH-9BA7DA/2020|EPI_ISL_573827|2</td><td>····················</td><td>····················</td><td>·····················</td><td>····················</td><td>····················</td><td>····················</td></tr>
<tr><td>L</td><td>7520</td><td>hCoV-19/Scotland/QEUH-9BA8D7/2020|EPI_ISL_573828|2</td><td>····················</td><td>····················</td><td>·····················</td><td>····················</td><td>····················</td><td>····················</td></tr>
<tr><td>L</td><td>7521</td><td>hCoV-19/Scotland/QEUH-9BA8E6/2020|EPI_ISL_573829|2</td><td>····················</td><td>····················</td><td>·····················</td><td>····················</td><td>····················</td><td>····················</td></tr>
<tr><td>L</td><td>7522</td><td>hCoV-19/Scotland/QEUH-9BA8F5/2020|EPI_ISL_573830|2</td><td>····················</td><td>····················</td><td>·····················</td><td>····················</td><td>····················</td><td>····················</td></tr>
<tr><td>L</td><td>7523</td><td>hCoV-19/Scotland/QEUH-9BA989/2020|EPI_ISL_573831|2</td><td>····················</td><td>····················</td><td>·····················</td><td>····················</td><td>····················</td><td>····················</td></tr>
<tr><td>L</td><td>7524</td><td>hCoV-19/Scotland/QEUH-9BAC44/2020|EPI_ISL_573836|2</td><td>····················</td><td>····················</td><td>·····················</td><td>····················</td><td>····················</td><td>····················</td></tr>
<tr><td>L</td><td>7525</td><td>hCoV-19/Scotland/QEUH-9BAD7E/2020|EPI_ISL_573837|2</td><td>····················</td><td>····················</td><td>·····················</td><td>····················</td><td>····················</td><td>····················</td></tr>
<tr><td>L</td><td>7526</td><td>hCoV-19/Scotland/QEUH-9BAD8D/2020|EPI_ISL_573838|2</td><td>····················</td><td>····················</td><td>·····················</td><td>····················</td><td>····················</td><td>····················</td></tr>
<tr><td>L</td><td>7527</td><td>hCoV-19/Scotland/QEUH-9BAD9C/2020|EPI_ISL_573839|2</td><td>····················</td><td>·A··················</td><td>·····················</td><td>····················</td><td>····················</td><td>··A·················</td></tr>
<tr><td>L</td><td>7528</td><td>hCoV-19/Scotland/QEUH-9BADBA/2020|EPI_ISL_573840|2</td><td>····················</td><td>····················</td><td>·····················</td><td>····················</td><td>····················</td><td>····················</td></tr>
<tr><td>L</td><td>7529</td><td>hCoV-19/Scotland/QEUH-9BADD8/2020|EPI_ISL_573841|2</td><td>····················</td><td>····················</td><td>·····················</td><td>····················</td><td>····················</td><td>····················</td></tr>
<tr><td>L</td><td>7530</td><td>hCoV-19/Scotland/QEUH-9BAE02/2020|EPI_ISL_573842|2</td><td>····················</td><td>····················</td><td>·····················</td><td>····················</td><td>····················</td><td>····················</td></tr>
<tr><td>L</td><td>7531</td><td>hCoV-19/Scotland/QEUH-9BAE4E/2020|EPI_ISL_573843|2</td><td>····················</td><td>····················</td><td>·····················</td><td>····················</td><td>····················</td><td>····················</td></tr>
<tr><td>L</td><td>7532</td><td>hCoV-19/Scotland/QEUH-9BAE5D/2020|EPI_ISL_573844|2</td><td>····················</td><td>····················</td><td>·····················</td><td>····················</td><td>····················</td><td>····················</td></tr>
<tr><td>L</td><td>7533</td><td>hCoV-19/Scotland/QEUH-9BAE6C/2020|EPI_ISL_573845|2</td><td>····················</td><td>····················</td><td>·····················</td><td>····················</td><td>····················</td><td>····················</td></tr>
<tr><td>L</td><td>7534</td><td>hCoV-19/Scotland/QEUH-9BAE7B/2020|EPI_ISL_573846|2</td><td>····················</td><td>····················</td><td>·····················</td><td>····················</td><td>····················</td><td>····················</td></tr>
<tr><td>L</td><td>7535</td><td>hCoV-19/Scotland/QEUH-9BAE8A/2020|EPI_ISL_573847|2</td><td>····················</td><td>····················</td><td>·····················</td><td>····················</td><td>····················</td><td>····················</td></tr>
<tr><td>L</td><td>7536</td><td>hCoV-19/Scotland/QEUH-9BAE99/2020|EPI_ISL_573848|2</td><td>····················</td><td>····················</td><td>·····················</td><td>····················</td><td>····················</td><td>····················</td></tr>
<tr><td>L</td><td>7537</td><td>hCoV-19/Scotland/QEUH-9BAEA8/2020|EPI_ISL_573849|2</td><td>····················</td><td>····················</td><td>·····················</td><td>····················</td><td>····················</td><td>····················</td></tr>
<tr><td>L</td><td>7538</td><td>hCoV-19/Wales/PHWC-16BF6F/2020|EPI_ISL_573993|2020</td><td>····················</td><td>····················</td><td>·····················</td><td>····················</td><td>····················</td><td>····················</td></tr>
<tr><td>L</td><td>7539</td><td>hCoV-19/Wales/PHWC-16BF7E/2020|EPI_ISL_573994|2020</td><td>····················</td><td>····················</td><td>·····················</td><td>····················</td><td>····················</td><td>····················</td></tr>
<tr><td>L</td><td>7540</td><td>hCoV-19/Wales/PHWC-16BF8D/2020|EPI_ISL_573995|2020</td><td>····················</td><td>····················</td><td>·····················</td><td>····················</td><td>····················</td><td>····················</td></tr>
<tr><td>L</td><td>7541</td><td>hCoV-19/Wales/PHWC-16BF9C/2020|EPI_ISL_573996|2020</td><td>····················</td><td>····················</td><td>·····················</td><td>····················</td><td>····················</td><td>····················</td></tr>
<tr><td>L</td><td>7542</td><td>hCoV-19/Wales/PHWC-16BFAB/2020|EPI_ISL_573997|2020</td><td>····················</td><td>····················</td><td>·····················</td><td>····················</td><td>····················</td><td>····················</td></tr>
<tr><td>L</td><td>7543</td><td>hCoV-19/Wales/PHWC-16BFBA/2020|EPI_ISL_573998|2020</td><td>····················</td><td>····················</td><td>·····················</td><td>····················</td><td>····················</td><td>····················</td></tr>
<tr><td>L</td><td>7544</td><td>hCoV-19/Wales/PHWC-16BFD8/2020|EPI_ISL_573999|2020</td><td>····················</td><td>····················</td><td>·····················</td><td>····················</td><td>····················</td><td>····················</td></tr>
</table>

| | | | | | | | |
|---|---|---|---|---|---|---|---|
| L | 7545 hCoV-19/Scotland/QEUH-9C9A0C/2020\|EPI_ISL_576071\|2 | ···················· | ···················· | ···················· | ···················· | ···················· | ···················· |
| L | 7546 hCoV-19/Scotland/QEUH-9C92D7/2020\|EPI_ISL_576072\|2 | ···················· | ···················· | ···················· | ···················· | ···················· | ···················· |
| L | 7547 hCoV-19/Scotland/QEUH-9C9CF7/2020\|EPI_ISL_576073\|2 | ···················· | ···················· | ···················· | ···················· | ···················· | ···················· |
| L | 7548 hCoV-19/Scotland/QEUH-9C9AA2/2020\|EPI_ISL_576074\|2 | ···················· | ···················· | ···················· | ···················· | ···················· | ···················· |
| L | 7549 hCoV-19/Scotland/QEUH-9CA467/2020\|EPI_ISL_576075\|2 | ···················· | ···················· | ···················· | ···················· | ···················· | ···················· |
| L | 7550 hCoV-19/Scotland/QEUH-9CA607/2020\|EPI_ISL_576076\|2 | ···················· | ···················· | ···················· | ···················· | ···················· | ···················· |
| L | 7551 hCoV-19/Scotland/QEUH-9C927D/2020\|EPI_ISL_576077\|2 | ···················· | ···················· | ···················· | ···················· | ···················· | ···················· |
| L | 7552 hCoV-19/Scotland/QEUH-9C9B63/2020\|EPI_ISL_576078\|2 | ···················· | ···················· | ···················· | ···················· | ···················· | ···················· |
| L | 7553 hCoV-19/Scotland/QEUH-9C92AA/2020\|EPI_ISL_576079\|2 | ···················· | ···················· | ···················· | ···················· | ···················· | ···················· |
| L | 7554 hCoV-19/Scotland/QEUH-9CA2C7/2020\|EPI_ISL_576080\|2 | ···················· | ···················· | ···················· | ···················· | ···················· | ···················· |
| L | 7555 hCoV-19/Scotland/QEUH-9CA1F7/2020\|EPI_ISL_576082\|2 | ···················· | ···················· | ···················· | ···················· | ···················· | ···················· |
| L | 7556 hCoV-19/Scotland/QEUH-9C9D03/2020\|EPI_ISL_576083\|2 | ···················· | ···················· | ···················· | ···················· | ···················· | ···················· |
| L | 7557 hCoV-19/Scotland/QEUH-9C9240/2020\|EPI_ISL_576084\|2 | ···················· | ···················· | ···················· | ···················· | ···················· | ···················· |
| L | 7558 hCoV-19/Scotland/QEUH-9CA34C/2020\|EPI_ISL_576085\|2 | ···················· | ···················· | ···················· | ···················· | ···················· | ···················· |
| L | 7559 hCoV-19/Scotland/QEUH-9C9653/2020\|EPI_ISL_576086\|2 | ···················· | ···················· | ···················· | ···················· | ···················· | ···················· |
| L | 7560 hCoV-19/Scotland/QEUH-9C9680/2020\|EPI_ISL_576087\|2 | ···················· | ···················· | ···················· | ···················· | ···················· | ···················· |
| L | 7561 hCoV-19/Scotland/QEUH-9C92B9/2020\|EPI_ISL_576089\|2 | ···················· | ···················· | ···················· | ···················· | ···················· | ···················· |
| L | 7562 hCoV-19/Scotland/QEUH-9C99E1/2020\|EPI_ISL_576090\|2 | ···················· | ···················· | ···················· | ···················· | ···················· | ···················· |
| L | 7563 hCoV-19/Scotland/QEUH-9C9D12/2020\|EPI_ISL_576091\|2 | ···················· | ···················· | ···················· | ···················· | ···················· | ···················· |
| L | 7564 hCoV-19/Scotland/QEUH-9C9B54/2020\|EPI_ISL_576092\|2 | ···················· | ···················· | ···················· | ···················· | ···················· | ···················· |
| L | 7565 hCoV-19/Scotland/QEUH-9C9C8E/2020\|EPI_ISL_576093\|2 | ···················· | ···················· | ···················· | ···················· | ···················· | ···················· |
| L | 7566 hCoV-19/England/MILK-9A67FF/2020\|EPI_ISL_576094\|20 | ···················· | ···················· | ···················· | ···················· | ···················· | ···················· |
| L | 7567 hCoV-19/USA/MI-MDHHS-SC21976/2020\|EPI_ISL_576208\|2 | ···················· | ···················· | ···················· | ···················· | ···················· | ···················· |
| L | 7568 hCoV-19/USA/MI-MDHHS-SC21986/2020\|EPI_ISL_576209\|2 | ···················· | ···················· | ···················· | ···················· | ···················· | ···················· |
| L | 7569 hCoV-19/Wales/PHWC-16DDCD/2020\|EPI_ISL_577535\|2020 | ···················· | ···················· | ···················· | ···················· | ···················· | ···················· |
| L | 7571 hCoV-19/USA/LA-EVTL462/2020\|EPI_ISL_578866\|2020-06 | ···················· | ···················· | ···················· | ···················· | ···················· | ···················· |
| L | 7572 hCoV-19/USA/LA-EVTL463/2020\|EPI_ISL_578867\|2020-06 | ···················· | ···················· | ···················· | ···················· | ···················· | ···················· |
| L | 7573 hCoV-19/USA/LA-EVTL464/2020\|EPI_ISL_578868\|2020-06 | ···················· | ···················· | ···················· | ···················· | ···················· | ···················· |
| L | 7574 hCoV-19/USA/LA-EVTL465/2020\|EPI_ISL_578869\|2020-06 | ···················· | ···················· | ···················· | ···················· | ···················· | ···················· |
| L | 7577 hCoV-19/New Zealand/20CV0398/2020\|EPI_ISL_582017\|2 | ···················· | ···················· | ···················· | ···················· | ···················· | ···················· |
| L | 7578 hCoV-19/New Zealand/20CV0401/2020\|EPI_ISL_582018\|2 | ···················· | ···················· | ···················· | ···················· | ···················· | ···················· |
| L | 7579 hCoV-19/New Zealand/20CV0408/2020\|EPI_ISL_582019\|2 | ···················· | ···················· | ···················· | ···················· | ···················· | ···················· |
| L | 7580 hCoV-19/New Zealand/20CV0409/2020\|EPI_ISL_582020\|2 | ···················· | ···················· | ···················· | ···················· | ···················· | ···················· |
| L | 7581 hCoV-19/New Zealand/20CV0414/2020\|EPI_ISL_582022\|2 | ···················· | ···················· | ···················· | ···················· | ···················· | ···················· |
| L | 7582 hCoV-19/New Zealand/20CV0415/2020\|EPI_ISL_582023\|2 | ···················· | ···················· | ···················· | ···················· | ···················· | ···················· |
| L | 7583 hCoV-19/Belgium/ULG-10460/2020\|EPI_ISL_582027\|2020 | ···················· | ···················· | ···················· | ···················· | ···················· | ···················· |
| L | 7584 hCoV-19/India/WB-IICB-030/2020\|EPI_ISL_582028\|2020 | ···················· | ···················· | ···················· | ···················· | ···················· | ···················· |
| L | 7585 hCoV-19/Iraq/ICGEB-5T/2020\|EPI_ISL_582030\|2020-06- | ···················· | ···················· | ···················· | ···················· | ···················· | ···················· |
| L | 7586 hCoV-19/Poland/IHG-PAS-8-29/2020\|EPI_ISL_582031\|20 | ···················· | ···················· | ···················· | ···················· | ···················· | ···················· |
| L | 7587 hCoV-19/Poland/IHG-PAS-8-30/2020\|EPI_ISL_582032\|20 | ···················· | ···················· | ···················· | ···················· | ···················· | ···················· |
| L | 7588 hCoV-19/Iran/1600/2020\|EPI_ISL_582033\|2020-04-03 | ···················· | ···················· | ···················· | ···················· | ···················· | ···················· |
| L | 7589 hCoV-19/Spain/PV-IBV-98007621/2020\|EPI_ISL_582048\| | ···················· | ···················· | ···················· | ···················· | ···················· | ···················· |
| L | 7590 hCoV-19/Spain/PV-IBV-98007637/2020\|EPI_ISL_582049\| | ···················· | ···················· | ···················· | ···················· | ···················· | ···················· |
| L | 7591 hCoV-19/Spain/PV-IBV-98007652/2020\|EPI_ISL_582050\| | ···················· | ···················· | ···················· | ···················· | ···················· | ···················· |
| L | 7592 hCoV-19/Spain/PV-IBV-98007662/2020\|EPI_ISL_582051\| | ···················· | ···················· | ···················· | ···················· | ···················· | ···················· |
| L | 7593 hCoV-19/Spain/PV-IBV-98007720/2020\|EPI_ISL_582052\| | ···················· | ···················· | ···················· | ···················· | ···················· | ···················· |
| L | 7594 hCoV-19/Spain/PV-IBV-98007364/2020\|EPI_ISL_582053\| | ···················· | ···················· | ···················· | ···················· | ···················· | ···················· |
| L | 7595 hCoV-19/Spain/PV-IBV-98007584/2020\|EPI_ISL_582054\| | ···················· | ···················· | ···················· | ···················· | ···················· | ···················· |
| L | 7596 hCoV-19/Spain/PV-IBV-98007592/2020\|EPI_ISL_582055\| | ···················· | ···················· | ···················· | ···················· | ···················· | ···················· |
| L | 7597 hCoV-19/Spain/PV-IBV-98007593/2020\|EPI_ISL_582056\| | ···················· | ···················· | ···················· | ···················· | ···················· | ···················· |
| L | 7598 hCoV-19/Spain/PV-IBV-98007600/2020\|EPI_ISL_582057\| | ···················· | ···················· | ···················· | ···················· | ···················· | ···················· |
| L | 7599 hCoV-19/Spain/PV-IBV-98007603/2020\|EPI_ISL_582058\| | ···················· | ···················· | ···················· | ···················· | ···················· | ···················· |
| L | 7600 hCoV-19/Spain/PV-IBV-98007609/2020\|EPI_ISL_582059\| | ···················· | ···················· | ···················· | ···················· | ···················· | ···················· |
| L | 7601 hCoV-19/Spain/PV-IBV-98007610/2020\|EPI_ISL_582060\| | ···················· | ···················· | ···················· | ···················· | ···················· | ···················· |
| L | 7602 hCoV-19/Spain/PV-IBV-98007612/2020\|EPI_ISL_582061\| | ···················· | ···················· | ···················· | ···················· | ···················· | ···················· |
| L | 7603 hCoV-19/Spain/PV-IBV-98007620/2020\|EPI_ISL_582062\| | ···················· | ···················· | ···················· | ···················· | ···················· | ···················· |
| L | 7604 hCoV-19/Spain/PV-IBV-98007622/2020\|EPI_ISL_582063\| | ···················· | ···················· | ···················· | ···················· | ···················· | ···············A···· |
| L | 7605 hCoV-19/Spain/PV-IBV-98007625/2020\|EPI_ISL_582064\| | ···················· | ···················· | ···················· | ···················· | ···················· | ···················· |
| L | 7606 hCoV-19/Spain/PV-IBV-98007627/2020\|EPI_ISL_582065\| | ···················· | ···················· | ···················· | ···················· | ···················· | ···················· |
| L | 7607 hCoV-19/Spain/PV-IBV-98007628/2020\|EPI_ISL_582066\| | ···················· | ···················· | ···················· | ···················· | ···················· | ···················· |
| L | 7608 hCoV-19/USA/MI-UM-10035812504/2020\|EPI_ISL_583310\| | ···················· | ·A·················· | ···················· | ···················· | ···················· | ···················· |
| L | 7609 hCoV-19/USA/MI-UM-10035748873/2020\|EPI_ISL_583312\| | ···················· | ···················· | ···················· | ···················· | ···················· | ···················· |
| L | 7610 hCoV-19/USA/MI-UM-10035752970/2020\|EPI_ISL_583313\| | ···················· | ·A·················· | ···················· | ···················· | ···················· | ···················· |
| L | 7611 hCoV-19/USA/MI-UM-10035736384/2020\|EPI_ISL_583314\| | ···················· | ···················· | ···················· | ···················· | ···················· | ···················· |
| L | 7612 hCoV-19/USA/MI-UM-10035725747/2020\|EPI_ISL_583315\| | ···················· | ·············A······ | ···················· | ···················· | ···················· | ···················· |
| L | 7613 hCoV-19/USA/MI-UM-10035790107/2020\|EPI_ISL_583316\| | ···················· | ···················· | ···················· | ···················· | ···················· | ···················· |
| L | 7614 hCoV-19/USA/MI-UM-10035803894/2020\|EPI_ISL_583317\| | ···················· | ···················· | ···················· | ···················· | ···················· | ···················· |
| L | 7615 hCoV-19/USA/MI-UM-10035822253/2020\|EPI_ISL_583318\| | ···················· | ·A·················· | ···················· | ···················· | ···················· | ···················· |
| L | 7616 hCoV-19/USA/MI-UM-10035790102/2020\|EPI_ISL_583319\| | ···················· | ···················· | ···················· | ···················· | ···················· | ···················· |
| L | 7617 hCoV-19/USA/MI-UM-10035813057/2020\|EPI_ISL_583320\| | ···················· | ·············A······ | ···················· | ···················· | ···················· | ···················· |
| L | 7619 hCoV-19/USA/MI-UM-10035812168/2020\|EPI_ISL_583324\| | ···················· | ···················· | ···················· | ···················· | ···················· | ···················· |
| L | 7620 hCoV-19/USA/MI-UM-10035768201/2020\|EPI_ISL_583325\| | ···················· | ·············A······ | ···················· | ···················· | ···················· | ···················· |
| L | 7621 hCoV-19/USA/MI-UM-10035804797/2020\|EPI_ISL_583326\| | ···················· | ···················· | ···················· | ···················· | ···················· | ···················· |
| L | 7622 hCoV-19/USA/MI-UM-10035806702/2020\|EPI_ISL_583327\| | T··················· | ·A·················· | ···················· | ···················· | ···················· | ···················· |
| L | 7624 hCoV-19/USA/MI-UM-10035889322/2020\|EPI_ISL_583329\| | T··················· | ·A·················· | ···················· | ···················· | ···················· | ···················· |
| L | 7626 hCoV-19/USA/MI-UM-10035886189/2020\|EPI_ISL_583331\| | ···················· | ·A·················· | ···················· | ···················· | ···················· | ···················· |
| L | 7627 hCoV-19/USA/MI-UM-10035854187/2020\|EPI_ISL_583332\| | ···················· | ···················· | ···················· | ···················· | ···················· | ···················· |
| L | 7631 hCoV-19/USA/MI-UM-10035849697/2020\|EPI_ISL_583336\| | T··················· | ·A·················· | ···················· | ···················· | ···················· | ···················· |
| L | 7632 hCoV-19/USA/MI-UM-10035889297/2020\|EPI_ISL_583337\| | T··················· | ·A·················· | ···················· | ···················· | ···················· | ···················· |
| L | 7633 hCoV-19/USA/MI-UM-10035869177/2020\|EPI_ISL_583340\| | ···················· | ···················· | ···················· | ···················· | ···················· | ···················· |

| | | | | | | | | |
|---|---|---|---|---|---|---|---|---|
| L | 7634 | hCoV-19/USA/MI-UM-10035843632/2020\|EPI_ISL_583341\| | ···················· | ···················· | ···················· | ···················· | ···················· | ···················· |
| L | 7635 | hCoV-19/USA/MI-UM-10035836175/2020\|EPI_ISL_583342\| | T··················· | ·A·················· | ···················· | ···················· | ···················· | ···················· |
| L | 7636 | hCoV-19/USA/MI-UM-10035863299/2020\|EPI_ISL_583343\| | T··················· | ·A·················· | ···················· | ···················· | ···················· | ···················· |
| L | 7637 | hCoV-19/USA/MI-UM-10035890097/2020\|EPI_ISL_583344\| | ···················· | ···················· | ···················· | ···················· | ···················· | ···················· |
| L | 7638 | hCoV-19/USA/MI-UM-10035851052/2020\|EPI_ISL_583345\| | ···················· | ···················· | ···················· | ···················· | ···················· | ···················· |
| L | 7641 | hCoV-19/USA/MI-UM-10035989687/2020\|EPI_ISL_583349\| | ···················· | ···················· | ···················· | ···················· | ···················· | ···················· |
| L | 7642 | hCoV-19/USA/MI-UM-10035956870/2020\|EPI_ISL_583350\| | ···················· | ···················· | ···················· | ···················· | ···················· | ···················· |
| L | 7643 | hCoV-19/USA/MI-UM-10035956808/2020\|EPI_ISL_583351\| | ···················· | ···················· | ···················· | ···················· | ···················· | ···················· |
| L | 7644 | hCoV-19/USA/MI-UM-10035980880/2020\|EPI_ISL_583352\| | ···················· | ···················· | ···················· | ···················· | ···················· | ···················· |
| L | 7645 | hCoV-19/USA/MI-UM-10035955134/2020\|EPI_ISL_583353\| | ···················· | ···················· | ···················· | ···················· | ···················· | ···················· |
| L | 7646 | hCoV-19/USA/MI-UM-10035952713/2020\|EPI_ISL_583354\| | ···················· | ···················· | ···················· | ···················· | ···················· | ···················· |
| L | 7647 | hCoV-19/USA/MI-UM-10035977253/2020\|EPI_ISL_583356\| | ···················· | ···················· | ···················· | ···················· | ···················· | ···················· |
| L | 7650 | hCoV-19/USA/MI-UM-10036024704/2020\|EPI_ISL_583362\| | ···················· | ···················· | ···················· | ···················· | ···················· | ···················· |
| L | 7652 | hCoV-19/USA/MI-UM-10035987523/2020\|EPI_ISL_583364\| | ···················· | ·A···A·············· | ···················· | ···················· | ···················· | ···················· |
| L | 7653 | hCoV-19/USA/MI-UM-10035986324/2020\|EPI_ISL_583365\| | ···················· | ···················· | ···················· | ···················· | ···················· | ···················· |
| L | 7655 | hCoV-19/USA/MI-UM-10035981014/2020\|EPI_ISL_583368\| | ···················· | ·A·················· | ···················· | ···················· | ···················· | ···················· |
| L | 7656 | hCoV-19/USA/MI-UM-10035963882/2020\|EPI_ISL_583369\| | ···················· | ···················· | ···················· | ···················· | ···················· | ···················· |
| L | 7661 | hCoV-19/USA/MI-UM-10036033491/2020\|EPI_ISL_583376\| | ···················· | ···················· | ···················· | ···················· | ···················· | ···················· |
| L | 7664 | hCoV-19/USA/MI-UM-10036036877/2020\|EPI_ISL_583379\| | ···················· | ·A·················· | ···················· | ···················· | ···················· | ···A················ |
| L | 7665 | hCoV-19/USA/MI-UM-10036021679/2020\|EPI_ISL_583380\| | ···················· | ···················· | ···················· | ···················· | ···················· | ···················· |
| L | 7666 | hCoV-19/USA/MI-UM-10036024406/2020\|EPI_ISL_583381\| | ···················· | ···················· | ···················· | ···················· | ···················· | ···················· |
| L | 7668 | hCoV-19/USA/MI-UM-10036005973/2020\|EPI_ISL_583384\| | ···················· | ···················· | ···················· | ···················· | ···················· | ···················· |
| L | 7670 | hCoV-19/USA/MI-UM-10035894711/2020\|EPI_ISL_583386\| | ···················· | ···················· | ···················· | ···················· | ···················· | ···················· |
| L | 7671 | hCoV-19/USA/MI-UM-10035923823/2020\|EPI_ISL_583387\| | T··················· | ·A·················· | ···················· | ···················· | ···················· | ···················· |
| L | 7674 | hCoV-19/USA/MI-UM-10035893881/2020\|EPI_ISL_583391\| | ···················· | ·A·················· | ···················· | ···················· | ···················· | ···················· |
| L | 7675 | hCoV-19/USA/MI-UM-10035907994/2020\|EPI_ISL_583392\| | ···················· | ···················· | ···················· | ···················· | ···················· | ···················· |
| L | 7676 | hCoV-19/USA/MI-UM-10035923573/2020\|EPI_ISL_583393\| | ···················· | ···················· | ···················· | ···················· | ···················· | ···················· |
| L | 7677 | hCoV-19/USA/MI-UM-10035926031/2020\|EPI_ISL_583394\| | ···················· | ·A·················· | ···················· | ···················· | ···················· | ···················· |
| L | 7678 | hCoV-19/USA/MI-UM-10035926038/2020\|EPI_ISL_583395\| | ···················· | ·A·················· | ···················· | ···················· | ···················· | ···················· |
| L | 7679 | hCoV-19/USA/MI-UM-10035892209/2020\|EPI_ISL_583396\| | ···················· | ·A·················· | ···················· | ···················· | ···················· | ···················· |
| L | 7680 | hCoV-19/USA/MI-UM-10035908071/2020\|EPI_ISL_583397\| | ···················· | ·A·················· | ···················· | ···················· | ···················· | ···················· |
| L | 7681 | hCoV-19/USA/MI-QDX-465/2020\|EPI_ISL_571200\|2020-03 | ···················· | ···················· | ···················· | ···················· | ···················· | ···················· |
| L | 7682 | hCoV-19/USA/MI-QDX-466/2020\|EPI_ISL_571201\|2020-03 | ···················· | ···················· | ···················· | ···················· | ···················· | ···················· |
| L | 7683 | hCoV-19/USA/MI-QDX-467/2020\|EPI_ISL_571202\|2020-03 | ···················· | ···················· | ···················· | ···················· | ···················· | ···················· |
| L | 7684 | hCoV-19/USA/MI-QDX-468/2020\|EPI_ISL_571203\|2020-03 | ···················· | ···················· | ···················· | ···················· | ···················· | ···················· |
| L | 7685 | hCoV-19/USA/MI-QDX-469/2020\|EPI_ISL_571204\|2020-03 | ···················· | ···················· | ···················· | ···················· | ···················· | ···················· |
| L | 7686 | hCoV-19/USA/MI-QDX-471/2020\|EPI_ISL_571205\|2020-03 | ···················· | ···················· | ···················· | ···················· | ···················· | ···················· |
| L | 7687 | hCoV-19/USA/MI-QDX-472/2020\|EPI_ISL_571206\|2020-03 | ···················· | ···················· | ···················· | ···················· | ···················· | ···················· |
| L | 7688 | hCoV-19/USA/MI-QDX-473/2020\|EPI_ISL_571207\|2020-03 | ···················· | ···················· | ···················· | ···················· | ···················· | ···················· |
| L | 7689 | hCoV-19/USA/MI-QDX-474/2020\|EPI_ISL_571208\|2020-03 | ···················· | ···················· | ···················· | ···················· | ···················· | ···················· |
| L | 7690 | hCoV-19/USA/MI-QDX-475/2020\|EPI_ISL_571209\|2020-03 | ···················· | ···················· | ···················· | ···················· | ···················· | ···················· |
| L | 7691 | hCoV-19/USA/ID-QDX-604/2020\|EPI_ISL_571299\|2020-03 | ···················· | ···················· | ···················· | ···················· | ···················· | ···················· |
| L | 7692 | hCoV-19/Scotland/QEUH-9BAA86/2020\|EPI_ISL_573832\|2 | ···················· | ···················· | ···················· | ···················· | ···················· | ···················· |
| L | 7693 | hCoV-19/Scotland/QEUH-9BAB38/2020\|EPI_ISL_573833\|2 | ···················· | ···················· | ···················· | ···················· | ···················· | ···················· |
| L | 7694 | hCoV-19/Scotland/QEUH-9BAC26/2020\|EPI_ISL_573834\|2 | ···················· | ···················· | ···················· | ···················· | ···················· | ···················· |
| L | 7695 | hCoV-19/Scotland/QEUH-9BAC35/2020\|EPI_ISL_573835\|2 | ···················· | ···················· | ···················· | ···················· | ···················· | ···················· |
| L | 7696 | hCoV-19/Scotland/QEUH-9BAEE4/2020\|EPI_ISL_573850\|2 | ···················· | ···················· | ···················· | ···················· | ···················· | ···················· |
| L | 7697 | hCoV-19/Scotland/QEUH-9BAEF3/2020\|EPI_ISL_573851\|2 | ···················· | ···················· | ···················· | ···················· | ···················· | ···················· |
| L | 7698 | hCoV-19/Scotland/QEUH-9BAF0F/2020\|EPI_ISL_573852\|2 | ···················· | ···················· | ···················· | ···················· | ···················· | ···················· |
| L | 7699 | hCoV-19/Scotland/QEUH-9BAFD2/2020\|EPI_ISL_573853\|2 | ···················· | ···················· | ···················· | ···················· | ···················· | ···················· |
| L | 7700 | hCoV-19/Scotland/QEUH-9BAFE1/2020\|EPI_ISL_573854\|2 | ···················· | ·A·················· | ···················· | ···················· | ···················· | ···A················ |
| L | 7701 | hCoV-19/Scotland/QEUH-9BAFF0/2020\|EPI_ISL_573855\|2 | ···················· | ···················· | ···················· | ···················· | ···················· | ···················· |
| L | 7702 | hCoV-19/Scotland/QEUH-9D1597/2020\|EPI_ISL_573856\|2 | ···················· | ···················· | ···················· | ···················· | ···················· | ···················· |
| L | 7703 | hCoV-19/Scotland/QEUH-9D1719/2020\|EPI_ISL_573857\|2 | ···················· | ···················· | ···················· | ···················· | ···················· | ···················· |
| L | 7704 | hCoV-19/Scotland/QEUH-9D1755/2020\|EPI_ISL_573858\|2 | ···················· | ···················· | ···················· | ···················· | ···················· | ···················· |
| L | 7705 | hCoV-19/Scotland/QEUH-9D195F/2020\|EPI_ISL_573859\|2 | ···················· | ···················· | ···················· | ···················· | ···················· | ···················· |
| L | 7706 | hCoV-19/Scotland/QEUH-9D196E/2020\|EPI_ISL_573860\|2 | ···················· | ···················· | ···················· | ···················· | ···················· | ···················· |
| L | 7707 | hCoV-19/Scotland/QEUH-9D1A89/2020\|EPI_ISL_573861\|2 | ···················· | ···················· | ···················· | ···················· | ···················· | ···················· |
| L | 7708 | hCoV-19/Scotland/QEUH-9D1AD4/2020\|EPI_ISL_573862\|2 | ···················· | ···················· | ···················· | ···················· | ···················· | ···················· |
| L | 7709 | hCoV-19/Scotland/QEUH-9D1AE3/2020\|EPI_ISL_573863\|2 | ···················· | ···················· | ···················· | ···················· | ···················· | ···················· |
| L | 7710 | hCoV-19/Scotland/QEUH-9D1BB3/2020\|EPI_ISL_573864\|2 | ···················· | ···················· | ···················· | ···················· | ···················· | ···················· |
| L | 7711 | hCoV-19/Scotland/QEUH-9D1C1A/2020\|EPI_ISL_573865\|2 | ···················· | ···················· | ···················· | ···················· | ···················· | ···················· |
| L | 7712 | hCoV-19/Wales/ALDP-9BCED3/2020\|EPI_ISL_573866\|2020 | ···················· | ···················· | ···················· | ···················· | ···················· | ···················· |
| L | 7713 | hCoV-19/Wales/CAMC-9B1E1A/2020\|EPI_ISL_573867\|2020 | ···················· | ···················· | ···················· | ···················· | ···················· | ···················· |
| L | 7714 | hCoV-19/Wales/CAMC-9C00B8/2020\|EPI_ISL_573868\|2020 | ···················· | ···················· | ···················· | ···················· | ···················· | ···················· |
| L | 7715 | hCoV-19/Wales/CAMC-9C1B6B/2020\|EPI_ISL_573869\|2020 | ···················· | ···················· | ···················· | ···················· | ···················· | ···················· |
| L | 7717 | hCoV-19/Wales/MILK-9C729D/2020\|EPI_ISL_573871\|2020 | ···················· | ···················· | ···················· | ···················· | ···················· | ···················· |
| L | 7718 | hCoV-19/Scotland/QEUH-9C87E8/2020\|EPI_ISL_576000\|2 | ···················· | ···················· | ···················· | ···················· | ···················· | ···················· |
| L | 7719 | hCoV-19/Scotland/QEUH-9C7C80/2020\|EPI_ISL_576002\|2 | ···················· | ···················· | ···················· | ···················· | ···················· | ···················· |
| L | 7720 | hCoV-19/Scotland/QEUH-9C87F7/2020\|EPI_ISL_576006\|2 | ···················· | ···················· | ···················· | ···················· | ···················· | ···················· |
| L | 7721 | hCoV-19/Scotland/QEUH-9C8BBF/2020\|EPI_ISL_576007\|2 | ···················· | ···················· | ···················· | ···················· | ···················· | ···················· |
| L | 7722 | hCoV-19/Scotland/QEUH-9C8469/2020\|EPI_ISL_576008\|2 | ···················· | ···················· | ···················· | ···················· | ···················· | ···················· |
| L | 7723 | hCoV-19/Scotland/QEUH-9C8733/2020\|EPI_ISL_576009\|2 | ···················· | ···················· | ···················· | ···················· | ···················· | ···················· |
| L | 7724 | hCoV-19/Scotland/QEUH-9C8821/2020\|EPI_ISL_576010\|2 | ···················· | ···················· | ···················· | ···················· | ···················· | ···················· |
| L | 7725 | hCoV-19/Scotland/QEUH-9C9073/2020\|EPI_ISL_576011\|2 | ···················· | ···················· | ···················· | ···················· | ···················· | ···················· |
| L | 7726 | hCoV-19/Scotland/QEUH-9C7D14/2020\|EPI_ISL_576012\|2 | ···················· | ···················· | ···················· | ···················· | ···················· | ···················· |
| L | 7727 | hCoV-19/Scotland/QEUH-9C8F1D/2020\|EPI_ISL_576013\|2 | ···················· | ···················· | ···················· | ···················· | ···················· | ···················· |
| L | 7728 | hCoV-19/Scotland/QEUH-9C7CF9/2020\|EPI_ISL_576014\|2 | ···················· | ···················· | ···················· | ···················· | ···················· | ···················· |
| L | 7729 | hCoV-19/Scotland/QEUH-9C8EC5/2020\|EPI_ISL_576015\|2 | ···················· | ···················· | ···················· | ···················· | ···················· | ···················· |
| L | 7730 | hCoV-19/Scotland/QEUH-9C8056/2020\|EPI_ISL_576016\|2 | ···················· | ···················· | ···················· | ···················· | ···················· | ···················· |

| | | | | | | | |
|---|---|---|---|---|---|---|---|
| L | 7731 hCoV-19/Scotland/QEUH-9C8760/2020\|EPI_ISL_576017\|2 | ···················· | ···················· | ···················· | ···················· | ···················· | ···················· |
| L | 7732 hCoV-19/Scotland/QEUH-9C8E3E/2020\|EPI_ISL_576019\|2 | ···················· | ···················· | ···················· | ···················· | ···················· | ···················· |
| L | 7733 hCoV-19/Scotland/QEUH-9C7D32/2020\|EPI_ISL_576020\|2 | ···················· | ···················· | ···················· | ···················· | ···················· | ···················· |
| L | 7734 hCoV-19/Scotland/QEUH-9C7D41/2020\|EPI_ISL_576021\|2 | ···················· | ·A·················· | ···················· | ···················· | ···················· | ···················· |
| L | 7735 hCoV-19/Scotland/QEUH-9C8706/2020\|EPI_ISL_576022\|2 | ···················· | ···················· | ···················· | ···················· | ···················· | ···················· |
| L | 7736 hCoV-19/Scotland/QEUH-9C8E6B/2020\|EPI_ISL_576023\|2 | ···················· | ···················· | ···················· | ···················· | ···················· | ···················· |
| L | 7737 hCoV-19/Scotland/QEUH-9C8830/2020\|EPI_ISL_576024\|2 | ···················· | ···················· | ···················· | ···················· | ···················· | ···················· |
| L | 7738 hCoV-19/Scotland/QEUH-9C7D05/2020\|EPI_ISL_576025\|2 | ···················· | ···················· | ···················· | ···················· | ···················· | ···················· |
| L | 7739 hCoV-19/Scotland/QEUH-9C7CEA/2020\|EPI_ISL_576027\|2 | ···················· | ···················· | ···················· | ···················· | ···················· | ···················· |
| L | 7740 hCoV-19/Scotland/QEUH-9C8BCE/2020\|EPI_ISL_576028\|2 | ···················· | ···················· | ···················· | ···················· | ···················· | ···················· |
| L | 7741 hCoV-19/Scotland/QEUH-9C8742/2020\|EPI_ISL_576029\|2 | ···················· | ···················· | ···················· | ···················· | ···················· | ···················· |
| L | 7742 hCoV-19/Scotland/QEUH-9C8715/2020\|EPI_ISL_576030\|2 | ···················· | ···················· | ···················· | ···················· | ···················· | ···················· |
| L | 7743 hCoV-19/Scotland/QEUH-9C7D23/2020\|EPI_ISL_576031\|2 | ···················· | ···················· | ···················· | ···················· | ···················· | ···················· |
| L | 7744 hCoV-19/Scotland/QEUH-9C7CAE/2020\|EPI_ISL_576032\|2 | ···················· | ···················· | ···················· | ···················· | ···················· | ···················· |
| L | 7745 hCoV-19/Scotland/QEUH-9C8E5C/2020\|EPI_ISL_576033\|2 | ···················· | ···················· | ···················· | ···················· | ···················· | ···················· |
| L | 7746 hCoV-19/Scotland/QEUH-9C892E/2020\|EPI_ISL_576034\|2 | ···················· | ···················· | ···················· | ···················· | ···················· | ···················· |
| L | 7747 hCoV-19/Scotland/QEUH-9C879D/2020\|EPI_ISL_576035\|2 | ···················· | ···················· | ···················· | ···················· | ···················· | ···················· |
| L | 7748 hCoV-19/Scotland/QEUH-9C8803/2020\|EPI_ISL_576036\|2 | ···················· | ···················· | ···················· | ···················· | ···················· | ···················· |
| L | 7749 hCoV-19/Scotland/QEUH-9C7CBD/2020\|EPI_ISL_576037\|2 | ···················· | ···················· | ···················· | ···················· | ···················· | ···················· |
| L | 7750 hCoV-19/Scotland/QEUH-9C925F/2020\|EPI_ISL_576038\|2 | ···················· | ···················· | ···················· | ···················· | ···················· | ···················· |
| L | 7751 hCoV-19/Scotland/QEUH-9C8F3B/2020\|EPI_ISL_576039\|2 | ···················· | ···················· | ···················· | ···················· | ···················· | ···················· |
| L | 7752 hCoV-19/Scotland/QEUH-9C7CDB/2020\|EPI_ISL_576040\|2 | ···················· | ···················· | ···················· | ···················· | ···················· | ···················· |
| L | 7753 hCoV-19/Scotland/QEUH-9C8EB6/2020\|EPI_ISL_576041\|2 | ···················· | ···················· | ···················· | ···················· | ···················· | ···················· |
| L | 7754 hCoV-19/Scotland/QEUH-9C8751/2020\|EPI_ISL_576042\|2 | ···················· | ···················· | ···················· | ···················· | ···················· | ···················· |
| L | 7755 hCoV-19/Scotland/QEUH-9C7C9F/2020\|EPI_ISL_576043\|2 | ···················· | ···················· | ···················· | ···················· | ···················· | ···················· |
| L | 7756 hCoV-19/Scotland/QEUH-9C9B72/2020\|EPI_ISL_576045\|2 | ···················· | ···················· | ···················· | ···················· | ···················· | ···················· |
| L | 7757 hCoV-19/Scotland/QEUH-9C9A66/2020\|EPI_ISL_576046\|2 | ···················· | ···················· | ···················· | ···················· | ···················· | ···················· |
| L | 7758 hCoV-19/Scotland/QEUH-9C9D21/2020\|EPI_ISL_576047\|2 | ···················· | ···················· | ···················· | ···················· | ···················· | ···················· |
| L | 7759 hCoV-19/Scotland/QEUH-9C96BD/2020\|EPI_ISL_576048\|2 | ···················· | ···················· | ···················· | ···················· | ···················· | ···················· |
| L | 7760 hCoV-19/Scotland/QEUH-9C9CE8/2020\|EPI_ISL_576049\|2 | ···················· | ···················· | ···················· | ···················· | ···················· | ···················· |
| L | 7761 hCoV-19/Scotland/QEUH-9C9B81/2020\|EPI_ISL_576051\|2 | ···················· | ···················· | ···················· | ···················· | ···················· | ···················· |
| L | 7762 hCoV-19/Scotland/QEUH-9CA42B/2020\|EPI_ISL_576052\|2 | ···················· | ···················· | ···················· | ···················· | ···················· | ···················· |
| L | 7763 hCoV-19/Scotland/QEUH-9CA3A6/2020\|EPI_ISL_576053\|2 | ···················· | ···················· | ···················· | ···················· | ···················· | ···················· |
| L | 7764 hCoV-19/Scotland/QEUH-9C928C/2020\|EPI_ISL_576054\|2 | ···················· | ···················· | ···················· | ···················· | ···················· | ···················· |
| L | 7765 hCoV-19/Scotland/QEUH-9C9662/2020\|EPI_ISL_576055\|2 | ···················· | ···················· | ···················· | ···················· | ···················· | ···················· |
| L | 7766 hCoV-19/Scotland/QEUH-9CA379/2020\|EPI_ISL_576056\|2 | ···················· | ···················· | ···················· | ···················· | ···················· | ···················· |
| L | 7767 hCoV-19/Scotland/QEUH-9CA24F/2020\|EPI_ISL_576057\|2 | ···················· | ···················· | ···················· | ···················· | ···················· | ···················· |
| L | 7768 hCoV-19/Scotland/QEUH-9C9CAC/2020\|EPI_ISL_576058\|2 | ···················· | ···················· | ···················· | ···················· | ···················· | ···················· |
| L | 7769 hCoV-19/Scotland/QEUH-9CA3B5/2020\|EPI_ISL_576059\|2 | ···················· | ···················· | ···················· | ···················· | ···················· | ···················· |
| L | 7770 hCoV-19/Scotland/QEUH-9C96AE/2020\|EPI_ISL_576060\|2 | ···················· | ···················· | ···················· | ···················· | ···················· | ···················· |
| L | 7771 hCoV-19/Scotland/QEUH-9C929B/2020\|EPI_ISL_576061\|2 | ···················· | ···················· | ···················· | ···················· | ···················· | ···················· |
| L | 7772 hCoV-19/Scotland/QEUH-9C92C8/2020\|EPI_ISL_576062\|2 | ···················· | ···················· | ···················· | ···················· | ···················· | ···················· |
| L | 7773 hCoV-19/Scotland/QEUH-9CA26D/2020\|EPI_ISL_576063\|2 | ···················· | ···················· | ···················· | ···················· | ···················· | ···················· |
| L | 7774 hCoV-19/Scotland/QEUH-9C9978/2020\|EPI_ISL_576065\|2 | ···················· | ···················· | ···················· | ···················· | ···················· | ···················· |
| L | 7775 hCoV-19/Scotland/QEUH-9C9671/2020\|EPI_ISL_576066\|2 | ···················· | ···················· | ···················· | ···················· | ···················· | ···················· |
| L | 7776 hCoV-19/Scotland/QEUH-9CA4B2/2020\|EPI_ISL_576069\|2 | ···················· | ···················· | ···················· | ···················· | ···················· | ···················· |
| L | 7777 hCoV-19/England/QEUH-9D343E/2020\|EPI_ISL_577333\|20 | ···················· | ···················· | ···················· | ···················· | ···················· | ···················· |
| L | 7778 hCoV-19/England/QEUH-9D34D4/2020\|EPI_ISL_577335\|20 | ···················· | ···················· | ···················· | ···················· | ·····T·············· | ···················· |
| L | 7779 hCoV-19/England/QEUH-9D3595/2020\|EPI_ISL_577339\|20 | ···················· | ···················· | ···················· | ···················· | ···················· | ···················· |
| L | 7780 hCoV-19/England/QEUH-9D35A4/2020\|EPI_ISL_577340\|20 | ···················· | ···················· | ···················· | ···················· | ···················· | ···················· |
| L | 7786 hCoV-19/Scotland/QEUH-9AE557/2020\|EPI_ISL_577359\|2 | ···················· | ···················· | ···················· | ···················· | ···················· | ···················· |
| L | 7788 hCoV-19/Scotland/QEUH-9AE5DF/2020\|EPI_ISL_577361\|2 | ···················· | ···················· | ···················· | ···················· | ···················· | ···················· |
| L | 7789 hCoV-19/Scotland/QEUH-9AE654/2020\|EPI_ISL_577363\|2 | ···················· | ···················· | ···················· | ···················· | ···················· | ···················· |
| L | 7790 hCoV-19/Scotland/QEUH-9AE672/2020\|EPI_ISL_577364\|2 | ···················· | ···················· | ···················· | ···················· | ···················· | ···················· |
| L | 7791 hCoV-19/Scotland/QEUH-9AE681/2020\|EPI_ISL_577365\|2 | ···················· | ···················· | ···················· | ···················· | ···················· | ···················· |
| L | 7792 hCoV-19/Scotland/QEUH-9AE6BE/2020\|EPI_ISL_577366\|2 | ···················· | ···················· | ···················· | ···················· | ···················· | ···················· |
| L | 7793 hCoV-19/Scotland/QEUH-9AE706/2020\|EPI_ISL_577367\|2 | ···················· | ···················· | ···················· | ···················· | ···················· | ···················· |
| L | 7794 hCoV-19/Scotland/QEUH-9AE724/2020\|EPI_ISL_577369\|2 | ···················· | ···················· | ···················· | ···················· | ···················· | ···················· |
| L | 7795 hCoV-19/Scotland/QEUH-9AE751/2020\|EPI_ISL_577370\|2 | ···················· | ···················· | ···················· | ···················· | ···················· | ···················· |
| L | 7796 hCoV-19/Scotland/QEUH-9AE77F/2020\|EPI_ISL_577371\|2 | ···················· | ···················· | ···················· | ···················· | ···················· | ···················· |
| L | 7797 hCoV-19/Scotland/QEUH-9AE87C/2020\|EPI_ISL_577372\|2 | ···················· | ···················· | ···················· | ···················· | ···················· | ···················· |
| L | 7798 hCoV-19/Scotland/QEUH-9AE900/2020\|EPI_ISL_577373\|2 | ···················· | ···················· | ···················· | ···················· | ···················· | ···················· |
| L | 7799 hCoV-19/Scotland/QEUH-9D1078/2020\|EPI_ISL_577375\|2 | ···················· | ·A·················· | ···················· | ···················· | ···················· | ···················· |
| L | 7800 hCoV-19/Scotland/QEUH-9D1096/2020\|EPI_ISL_577376\|2 | ···················· | ···················· | ···················· | ···················· | ···T················ | ···················· |
| L | 7801 hCoV-19/Scotland/QEUH-9D1272/2020\|EPI_ISL_577377\|2 | ···················· | ···················· | ···················· | ···················· | ···················· | ···················· |
| L | 7802 hCoV-19/Scotland/QEUH-9D12CD/2020\|EPI_ISL_577378\|2 | ···················· | ···················· | ···················· | ···················· | ···················· | ···················· |
| L | 7803 hCoV-19/Scotland/QEUH-9D145E/2020\|EPI_ISL_577379\|2 | ···················· | ···················· | ···················· | ···················· | ···················· | ···················· |
| L | 7804 hCoV-19/Scotland/QEUH-9D15C4/2020\|EPI_ISL_577380\|2 | ···················· | ·A·················· | ···················· | ···················· | ···················· | ··A················· |
| L | 7805 hCoV-19/Scotland/QEUH-9D3252/2020\|EPI_ISL_577382\|2 | ···················· | ···················· | ···················· | ···················· | ···················· | ···················· |
| L | 7806 hCoV-19/Wales/ALDP-9D8BF8/2020\|EPI_ISL_577383\|2020 | ···················· | ···················· | ···················· | ···················· | ···················· | ···················· |
| L | 7807 hCoV-19/Wales/ALDP-9D9492/2020\|EPI_ISL_577384\|2020 | ···················· | ···················· | ···················· | ···················· | ···················· | ···················· |
| L | 7808 hCoV-19/Wales/MILK-9D52BA/2020\|EPI_ISL_577387\|2020 | ···················· | ···················· | ···················· | ···················· | ···················· | ···················· |
| L | 7816 hCoV-19/USA/WI-WSLH-200512/2020\|EPI_ISL_578653\|202 | ···················· | ···················· | ···················· | ···················· | ···················· | ···················· |
| L | 7817 hCoV-19/USA/WI-WSLH-200513/2020\|EPI_ISL_578654\|202 | ···················· | ···················· | ···················· | ···················· | ···················· | ···················· |
| L | 7820 hCoV-19/USA/WI-WSLH-200517/2020\|EPI_ISL_578657\|202 | ···················· | ···················· | ···················· | ···················· | ···················· | ···················· |
| L | 7821 hCoV-19/USA/WI-WSLH-200518/2020\|EPI_ISL_578658\|202 | ···················· | ···················· | ···················· | ···················· | ···················· | ···················· |
| L | 7822 hCoV-19/USA/WI-WSLH-200519/2020\|EPI_ISL_578659\|202 | ···················· | ···················· | ···················· | ···················· | ···················· | ···················· |
| L | 7823 hCoV-19/USA/WI-WSLH-200520/2020\|EPI_ISL_578660\|202 | ···················· | ···················· | ···················· | ···················· | ···················· | ···················· |
| L | 7824 hCoV-19/USA/WI-WSLH-200521/2020\|EPI_ISL_578661\|202 | ···················· | ···················· | ···················· | ···················· | ···················· | ···················· |
| L | 7825 hCoV-19/USA/WI-WSLH-200522/2020\|EPI_ISL_578662\|202 | ···················· | ···················· | ···················· | ···················· | ···················· | ···················· |

| | | | | | | | |
|---|---|---|---|---|---|---|---|
| L | 7826 hCoV-19/USA/WI-WSLH-200523/2020\|EPI_ISL_578663\|202 | ···················· | ···················· | ···················· | ···················· | ···················· | ···················· |
| L | 7827 hCoV-19/USA/WI-WSLH-200524/2020\|EPI_ISL_578664\|202 | ···················· | ···················· | ···················· | ···················· | ···················· | ···················· |
| L | 7828 hCoV-19/USA/WI-WSLH-200526/2020\|EPI_ISL_578665\|202 | ···················· | ···················· | ···················· | ···················· | ···················· | ···················· |
| L | 7829 hCoV-19/USA/WI-WSLH-200527/2020\|EPI_ISL_578666\|202 | ···················· | ···················· | ···················· | ···················· | ···················· | ···················· |
| L | 7830 hCoV-19/USA/WI-WSLH-200528/2020\|EPI_ISL_578667\|202 | ···················· | ···················· | ···················· | ···················· | ···················· | ···················· |
| L | 7831 hCoV-19/USA/WI-WSLH-200529/2020\|EPI_ISL_578668\|202 | ···················· | ···················· | ···················· | ···················· | ···················· | ···················· |
| L | 7832 hCoV-19/USA/WI-WSLH-200530/2020\|EPI_ISL_578669\|202 | ···················· | ···················· | ···················· | ···················· | ···················· | ···················· |
| L | 7833 hCoV-19/USA/WI-WSLH-200531/2020\|EPI_ISL_578670\|202 | ···················· | ···················· | ···················· | ···················· | ···················· | ···················· |
| L | 7834 hCoV-19/USA/WI-WSLH-200532/2020\|EPI_ISL_578671\|202 | ···················· | ···················· | ···················· | ···················· | ···················· | ···················· |
| L | 7835 hCoV-19/USA/WI-WSLH-200535/2020\|EPI_ISL_578672\|202 | ···················· | ···················· | ···················· | ···················· | ···················· | ···················· |
| L | 7836 hCoV-19/USA/WI-WSLH-200536/2020\|EPI_ISL_578673\|202 | ···················· | ···················· | ···················· | ···················· | ···················· | ···················· |
| L | 7837 hCoV-19/USA/WI-WSLH-200537/2020\|EPI_ISL_578674\|202 | ···················· | ···················· | ···················· | ···················· | ···················· | ···················· |
| L | 7838 hCoV-19/USA/WI-WSLH-200545/2020\|EPI_ISL_578675\|202 | ···················· | ···················· | ···················· | ···················· | ···················· | ···················· |
| L | 7844 hCoV-19/USA/WI-WSLH-200569/2020\|EPI_ISL_578681\|202 | ···················· | ···················· | ···················· | ···················· | ···················· | ···················· |
| L | 7855 hCoV-19/USA/WI-WSLH-200581/2020\|EPI_ISL_578692\|202 | ···················· | ···················· | ···················· | ···················· | ···················· | ···················· |
| L | 7859 hCoV-19/USA/LA-EVTL321/2020\|EPI_ISL_578698\|2020-05 | ···················· | ···················· | ···················· | ···················· | ···················· | ···················· |
| L | 7860 hCoV-19/USA/LA-EVTL437/2020\|EPI_ISL_578699\|2020-06 | ···················· | ···················· | ···················· | ···················· | ···················· | ···················· |
| L | 7861 hCoV-19/USA/LA-EVTL438/2020\|EPI_ISL_578700\|2020-06 | ···················· | ···················· | ···················· | ···················· | ···················· | ···················· |
| L | 7862 hCoV-19/USA/LA-EVTL305/2020\|EPI_ISL_578712\|2020-05 | ···················· | ···················· | ···················· | ···················· | ···················· | ···················· |
| L | 7863 hCoV-19/USA/LA-EVTL306/2020\|EPI_ISL_578713\|2020-05 | ···················· | ···················· | ···················· | ···················· | ···················· | ···················· |
| L | 7864 hCoV-19/USA/LA-EVTL307/2020\|EPI_ISL_578714\|2020-05 | ···················· | ···················· | ···················· | ···················· | ···················· | ···················· |
| L | 7865 hCoV-19/USA/LA-EVTL309/2020\|EPI_ISL_578716\|2020-05 | ···················· | ···················· | ···················· | ···················· | ···················· | ···················· |
| L | 7866 hCoV-19/USA/LA-EVTL310/2020\|EPI_ISL_578717\|2020-05 | ···················· | ···················· | ···················· | ···················· | ···················· | ···················· |
| L | 7867 hCoV-19/USA/LA-EVTL311/2020\|EPI_ISL_578718\|2020-05 | ···················· | ···················· | ···················· | ···················· | ···················· | ···················· |
| L | 7868 hCoV-19/USA/LA-EVTL312/2020\|EPI_ISL_578719\|2020-05 | ···················· | ···················· | ···················· | ···················· | ···················· | ···················· |
| L | 7869 hCoV-19/USA/LA-EVTL313/2020\|EPI_ISL_578720\|2020-05 | ···················· | ···················· | ···················· | ···················· | ···················· | ···················· |
| L | 7870 hCoV-19/USA/LA-EVTL314/2020\|EPI_ISL_578721\|2020-05 | ···················· | ···················· | ···················· | ···················· | ···················· | ···················· |
| L | 7871 hCoV-19/USA/LA-EVTL315/2020\|EPI_ISL_578722\|2020-05 | ···················· | ···················· | ···················· | ···················· | ···················· | ···················· |
| L | 7872 hCoV-19/England/ALDP-9EAE36/2020\|EPI_ISL_579977\|20 | ···················· | ···················· | ···················· | ···················· | ···················· | ···················· |
| L | 7873 hCoV-19/England/ALDP-9EADEE/2020\|EPI_ISL_579980\|20 | ···················· | ···················· | ···················· | ···················· | ···················· | ···················· |
| L | 7874 hCoV-19/England/ALDP-9EAD57/2020\|EPI_ISL_579982\|20 | ···················· | ···················· | ···················· | ···················· | ···················· | ···················· |
| L | 7875 hCoV-19/England/ALDP-9EA3DD/2020\|EPI_ISL_579983\|20 | ···················· | ···················· | ···················· | ···················· | ···················· | ···················· |
| L | 7876 hCoV-19/England/ALDP-9E7631/2020\|EPI_ISL_579984\|20 | ···················· | ···················· | ···················· | ···················· | ···················· | ···················· |
| L | 7877 hCoV-19/England/ALDP-9E9DC1/2020\|EPI_ISL_579985\|20 | ···················· | ···················· | ···················· | ···················· | ···················· | ···················· |
| L | 7878 hCoV-19/England/ALDP-9EAC5A/2020\|EPI_ISL_579987\|20 | ···················· | ···················· | ···················· | ···················· | ···················· | ···················· |
| L | 7879 hCoV-19/England/ALDP-9EA7E0/2020\|EPI_ISL_579989\|20 | ···················· | ···················· | ···················· | ···················· | ···················· | ···················· |
| L | 7880 hCoV-19/England/ALDP-9EA1C4/2020\|EPI_ISL_579990\|20 | ···················· | ···················· | ···················· | ···················· | ···················· | ···················· |
| L | 7881 hCoV-19/England/ALDP-9EA364/2020\|EPI_ISL_579991\|20 | ···················· | ···················· | ···················· | ···················· | ····T··············· | ···················· |
| L | 7882 hCoV-19/England/ALDP-9E9CA6/2020\|EPI_ISL_579992\|20 | ···················· | ···················· | ···················· | ···················· | ···················· | ···················· |
| L | 7883 hCoV-19/England/ALDP-9E9F52/2020\|EPI_ISL_579993\|20 | ···················· | ···················· | ···················· | ···················· | ···················· | ···················· |
| L | 7884 hCoV-19/England/ALDP-9EAB8A/2020\|EPI_ISL_579994\|20 | ···················· | ···················· | ···················· | ···················· | ···················· | ···················· |
| L | 7885 hCoV-19/England/ALDP-9EA6C5/2020\|EPI_ISL_579995\|20 | ···················· | ···················· | ···················· | ···················· | ···················· | ···················· |
| L | 7886 hCoV-19/England/ALDP-9E7A08/2020\|EPI_ISL_579996\|20 | ···················· | ···················· | ···················· | ···················· | ···················· | ···················· |
| L | 7887 hCoV-19/England/ALDP-9EADA2/2020\|EPI_ISL_579997\|20 | ···················· | ···················· | ···················· | ···················· | ···················· | ···················· |
| L | 7888 hCoV-19/England/ALDP-9E9ACA/2020\|EPI_ISL_579999\|20 | ···················· | ···················· | ···················· | ···················· | ···················· | ···················· |
| L | 7889 hCoV-19/England/QEUH-9DC70F/2020\|EPI_ISL_580960\|20 | ···················· | ···················· | ···················· | ···················· | ···················· | ···················· |
| L | 7890 hCoV-19/Wales/MILK-9E0DAC/2020\|EPI_ISL_580961\|2020 | ···················· | ···················· | ···················· | ···················· | ···················· | ···················· |
| L | 7891 hCoV-19/England/QEUH-9DC7E1/2020\|EPI_ISL_580962\|20 | ···················· | ···················· | ···················· | ···················· | ···················· | ···················· |
| L | 7892 hCoV-19/England/QEUH-9DC875/2020\|EPI_ISL_580963\|20 | ···················· | ···················· | ···················· | ···················· | ···················· | ···················· |
| L | 7893 hCoV-19/England/QEUH-9DAE66/2020\|EPI_ISL_580964\|20 | ···················· | ···················· | ···················· | ···················· | ···················· | ···················· |
| L | 7894 hCoV-19/England/QEUH-9DB09C/2020\|EPI_ISL_580965\|20 | ···················· | ···················· | ···················· | ···················· | ···················· | ···················· |
| L | 7895 hCoV-19/England/QEUH-9DC71E/2020\|EPI_ISL_580966\|20 | ···················· | ···················· | ···················· | ···················· | ···················· | ···················· |
| L | 7896 hCoV-19/England/QEUH-9DAB05/2020\|EPI_ISL_580967\|20 | ···················· | ···················· | ···················· | ···················· | ···················· | ···················· |
| L | 7897 hCoV-19/England/QEUH-9DC523/2020\|EPI_ISL_580968\|20 | ···················· | ···················· | ···················· | ···················· | ···················· | ···················· |
| L | 7898 hCoV-19/England/QEUH-9DCA42/2020\|EPI_ISL_580969\|20 | ···················· | ···················· | ···················· | ···················· | ······T············· | ···················· |
| L | 7899 hCoV-19/England/QEUH-9DCC4C/2020\|EPI_ISL_580970\|20 | ···················· | ···················· | ···················· | ···················· | ···················· | ···················· |
| L | 7900 hCoV-19/England/QEUH-9DC884/2020\|EPI_ISL_580971\|20 | ···················· | ···················· | ···················· | ···················· | ···················· | ···················· |
| L | 7901 hCoV-19/England/QEUH-9DC699/2020\|EPI_ISL_580972\|20 | ···················· | ···················· | ···················· | ···················· | ···················· | ···················· |
| L | 7902 hCoV-19/England/QEUH-9DCB30/2020\|EPI_ISL_580973\|20 | ···················· | ·A·················· | ···················· | ···················· | ···················· | ···A················ |
| L | 7903 hCoV-19/England/QEUH-9DCA33/2020\|EPI_ISL_580974\|20 | ···················· | ···················· | ···················· | ···················· | ···················· | ···················· |
| L | 7904 hCoV-19/England/QEUH-9DD337/2020\|EPI_ISL_580975\|20 | ···················· | ···················· | ···················· | ···················· | ···················· | ···················· |
| L | 7905 hCoV-19/England/MILK-9E0EE5/2020\|EPI_ISL_580976\|20 | ···················· | ···················· | ···················· | ···················· | ···················· | ···················· |
| L | 7906 hCoV-19/England/QEUH-9DCA51/2020\|EPI_ISL_580977\|20 | ···················· | ···················· | ···················· | ···················· | ···················· | ···················· |
| L | 7907 hCoV-19/Wales/MILK-9E128E/2020\|EPI_ISL_580979\|2020 | ···················· | ···················· | ···················· | ···················· | ···················· | ···················· |
| L | 7908 hCoV-19/Wales/MILK-9E0DF7/2020\|EPI_ISL_580980\|2020 | ···················· | ···················· | ···················· | ···················· | ···················· | ···················· |
| L | 7909 hCoV-19/England/QEUH-9DAF09/2020\|EPI_ISL_580981\|20 | ···················· | ·A·················· | ···················· | ···················· | ···················· | ···················· |
| L | 7910 hCoV-19/England/QEUH-9DC857/2020\|EPI_ISL_580982\|20 | ···················· | ···················· | ···················· | ···················· | ···················· | ···················· |
| L | 7911 hCoV-19/England/QEUH-9DCBC7/2020\|EPI_ISL_580983\|20 | ···················· | ···················· | ···················· | ···················· | ···················· | ···················· |
| L | 7912 hCoV-19/England/QEUH-9DC7F0/2020\|EPI_ISL_580984\|20 | ···················· | ·A·················· | ···················· | ···················· | ···················· | ···A················ |
| L | 7913 hCoV-19/England/QEUH-9DB07E/2020\|EPI_ISL_580985\|20 | ···················· | ···················· | ···················· | ···················· | ···················· | ···················· |
| L | 7914 hCoV-19/England/QEUH-9DC7D2/2020\|EPI_ISL_580986\|20 | ···················· | ···················· | ···················· | ···················· | ···················· | ···················· |
| L | 7915 hCoV-19/England/QEUH-9DCEFB/2020\|EPI_ISL_580987\|20 | ···················· | ···················· | ···················· | ···················· | ······T············· | ···················· |
| L | 7916 hCoV-19/England/QEUH-9DA938/2020\|EPI_ISL_580988\|20 | ···················· | ···················· | ···················· | ···················· | ···················· | ···················· |
| L | 7917 hCoV-19/England/QEUH-9DAC3F/2020\|EPI_ISL_580989\|20 | ···················· | ···················· | ···················· | ···················· | ···················· | ···················· |
| L | 7918 hCoV-19/Wales/MILK-9E0FF1/2020\|EPI_ISL_580990\|2020 | ···················· | ···················· | ···················· | ···················· | ···················· | ···················· |
| L | 7919 hCoV-19/England/MILK-9E138B/2020\|EPI_ISL_580991\|20 | ···················· | ···················· | ···················· | ···················· | ···················· | ···················· |
| L | 7920 hCoV-19/England/QEUH-9DCC3D/2020\|EPI_ISL_580992\|20 | ···················· | ···················· | ···················· | ···················· | ···················· | ···················· |
| L | 7921 hCoV-19/Wales/MILK-9E1233/2020\|EPI_ISL_580993\|2020 | ···················· | ···················· | ···················· | ···················· | ···················· | ···················· |
| L | 7922 hCoV-19/England/QEUH-9DC5BA/2020\|EPI_ISL_580994\|20 | ···················· | ·A·················· | ···················· | ···················· | ···················· | ···A················ |
| L | 7923 hCoV-19/England/MILK-9E139A/2020\|EPI_ISL_580995\|20 | ···················· | ···················· | ···················· | ···················· | ···················· | ···················· |

| | | | | | | | | |
|---|---|---|---|---|---|---|---|---|
| L | 7924 | hCoV-19/England/QEUH-9DAD2D/2020\|EPI_ISL_580997\|20 | ···················· | ···················· | ···················· | ···················· | ···················· | ···················· |
| L | 7925 | hCoV-19/England/MILK-9E0F6A/2020\|EPI_ISL_580998\|20 | ···················· | ·A·················· | ···················· | ···················· | ···················· | ···A················ |
| L | 7926 | hCoV-19/England/QEUH-9DC7B4/2020\|EPI_ISL_580999\|20 | ···················· | ···················· | ···················· | ···················· | ···················· | ···················· |
| L | 7927 | hCoV-19/USA/MI-UM-10035713817/2020\|EPI_ISL_583277\| | ···················· | ·A·················· | ···················· | ···················· | ···················· | ···················· |
| L | 7928 | hCoV-19/USA/MI-UM-10035713605/2020\|EPI_ISL_583278\| | ···················· | ·············A······ | ···················· | ···················· | ···················· | ···················· |
| L | 7929 | hCoV-19/USA/MI-UM-10035692627/2020\|EPI_ISL_583279\| | ···················· | ···················· | ···················· | ···················· | ···················· | ···················· |
| L | 7930 | hCoV-19/USA/MI-UM-10035711830/2020\|EPI_ISL_583280\| | ···················· | ·A·················· | ···················· | ···················· | ···················· | ···················· |
| L | 7931 | hCoV-19/USA/MI-UM-10035711872/2020\|EPI_ISL_583281\| | ···················· | ···················· | ···················· | ···················· | ···················· | ···················· |
| L | 7932 | hCoV-19/USA/MI-UM-10035703434/2020\|EPI_ISL_583282\| | ···················· | ·A·················· | ···················· | ···················· | ···················· | ···················· |
| L | 7933 | hCoV-19/USA/MI-UM-10035704378/2020\|EPI_ISL_583283\| | ···················· | ·A····A············· | ···················· | ···················· | ···················· | ···················· |
| L | 7934 | hCoV-19/USA/MI-UM-10035637067/2020\|EPI_ISL_583284\| | ···················· | ···················· | ···················· | ···················· | ···················· | ···················· |
| L | 7935 | hCoV-19/USA/MI-UM-10035641365/2020\|EPI_ISL_583285\| | ···················· | ·A·················· | ···················· | ···················· | ···················· | ···················· |
| L | 7937 | hCoV-19/USA/MI-QDX-476/2020\|EPI_ISL_571210\|2020-03 | ···················· | ···················· | ···················· | ···················· | ···················· | ···················· |
| L | 7938 | hCoV-19/USA/MI-QDX-477/2020\|EPI_ISL_571211\|2020-03 | ···················· | ···················· | ···················· | ···················· | ···················· | ···················· |
| L | 7939 | hCoV-19/USA/MI-QDX-478/2020\|EPI_ISL_571212\|2020-03 | ···················· | ···················· | ···················· | ···················· | ···················· | ···················· |
| L | 7940 | hCoV-19/USA/MI-QDX-479/2020\|EPI_ISL_571213\|2020-03 | ···················· | ···················· | ···················· | ···················· | ···················· | ···················· |
| L | 7941 | hCoV-19/USA/MI-QDX-480/2020\|EPI_ISL_571214\|2020-03 | ···················· | ···················· | ···················· | ···················· | ···················· | ···················· |
| L | 7942 | hCoV-19/USA/MI-QDX-481/2020\|EPI_ISL_571215\|2020-03 | ···················· | ···················· | ···················· | ···················· | ···················· | ···················· |
| L | 7943 | hCoV-19/USA/MI-QDX-482/2020\|EPI_ISL_571216\|2020-03 | ···················· | ···················· | ···················· | ···················· | ···················· | ···················· |
| L | 7944 | hCoV-19/USA/MI-QDX-483/2020\|EPI_ISL_571217\|2020-03 | ···················· | ···················· | ···················· | ···················· | ···················· | ···················· |
| L | 7945 | hCoV-19/USA/MI-QDX-484/2020\|EPI_ISL_571218\|2020-03 | ···················· | ···················· | ···················· | ···················· | ···················· | ···················· |
| L | 7946 | hCoV-19/USA/MI-QDX-485/2020\|EPI_ISL_571219\|2020-03 | ···················· | ···················· | ···················· | ···················· | ···················· | ···················· |
| L | 7947 | hCoV-19/USA/MI-QDX-486/2020\|EPI_ISL_571220\|2020-03 | ···················· | ···················· | ···················· | ···················· | ···················· | ···················· |
| L | 7948 | hCoV-19/USA/MI-QDX-487/2020\|EPI_ISL_571221\|2020-03 | ···················· | ···················· | ···················· | ···················· | ···················· | ···················· |
| L | 7949 | hCoV-19/USA/MI-QDX-488/2020\|EPI_ISL_571222\|2020-03 | ···················· | ···················· | ···················· | ···················· | ···················· | ···················· |
| L | 7950 | hCoV-19/USA/MI-QDX-489/2020\|EPI_ISL_571223\|2020-03 | ···················· | ···················· | ···················· | ···················· | ···················· | ···················· |
| L | 7951 | hCoV-19/USA/MI-QDX-490/2020\|EPI_ISL_571224\|2020-03 | ···················· | ···················· | ···················· | ···················· | ···················· | ···················· |
| L | 7952 | hCoV-19/USA/MI-QDX-491/2020\|EPI_ISL_571225\|2020-03 | ···················· | ···················· | ···················· | ···················· | ···················· | ···················· |
| L | 7953 | hCoV-19/USA/MI-QDX-492/2020\|EPI_ISL_571226\|2020-03 | ···················· | ···················· | ···················· | ···················· | ···················· | ···················· |
| L | 7954 | hCoV-19/USA/MI-QDX-493/2020\|EPI_ISL_571227\|2020-03 | ···················· | ···················· | ···················· | ···················· | ···················· | ···················· |
| L | 7955 | hCoV-19/USA/MI-QDX-494/2020\|EPI_ISL_571228\|2020-03 | ···················· | ···················· | ···················· | ···················· | ···················· | ···················· |
| L | 7956 | hCoV-19/USA/MI-QDX-495/2020\|EPI_ISL_571229\|2020-03 | ···················· | ···················· | ···················· | ···················· | ···················· | ···················· |
| L | 7957 | hCoV-19/USA/MI-QDX-496/2020\|EPI_ISL_571230\|2020-03 | ···················· | ···················· | ···················· | ···················· | ···················· | ···················· |
| L | 7958 | hCoV-19/USA/MI-QDX-497/2020\|EPI_ISL_571231\|2020-03 | ···················· | ···················· | ···················· | ···················· | ···················· | ···················· |
| L | 7959 | hCoV-19/USA/MI-QDX-498/2020\|EPI_ISL_571232\|2020-03 | ···················· | ···················· | ···················· | ···················· | ···················· | ···················· |
| L | 7960 | hCoV-19/USA/MI-QDX-499/2020\|EPI_ISL_571233\|2020-03 | ···················· | ···················· | ···················· | ···················· | ···················· | ···················· |
| L | 7961 | hCoV-19/USA/MI-QDX-500/2020\|EPI_ISL_571234\|2020-03 | ···················· | ···················· | ···················· | ···················· | ···················· | ···················· |
| L | 7962 | hCoV-19/USA/MI-QDX-501/2020\|EPI_ISL_571235\|2020-03 | ···················· | ···················· | ···················· | ···················· | ···················· | ···················· |
| L | 7963 | hCoV-19/USA/MI-QDX-502/2020\|EPI_ISL_571236\|2020-03 | ···················· | ···················· | ···················· | ···················· | ···················· | ···················· |
| L | 7964 | hCoV-19/USA/MI-QDX-503/2020\|EPI_ISL_571237\|2020-03 | ···················· | ···················· | ···················· | ···················· | ···················· | ···················· |
| L | 7965 | hCoV-19/USA/MI-QDX-504/2020\|EPI_ISL_571238\|2020-03 | ···················· | ···················· | ···················· | ···················· | ···················· | ···················· |
| L | 7966 | hCoV-19/USA/MI-QDX-505/2020\|EPI_ISL_571239\|2020-03 | ···················· | ···················· | ···················· | ···················· | ···················· | ···················· |
| L | 7967 | hCoV-19/USA/MI-QDX-506/2020\|EPI_ISL_571240\|2020-03 | ···················· | ···················· | ···················· | ···················· | ···················· | ···················· |
| L | 7968 | hCoV-19/USA/MI-QDX-507/2020\|EPI_ISL_571241\|2020-03 | ···················· | ···················· | ···················· | ···················· | ···················· | ···················· |
| L | 7969 | hCoV-19/USA/MI-QDX-508/2020\|EPI_ISL_571242\|2020-03 | ···················· | ···················· | ···················· | ···················· | ···················· | ···················· |
| L | 7970 | hCoV-19/USA/MI-QDX-509/2020\|EPI_ISL_571243\|2020-03 | ···················· | ···················· | ···················· | ···················· | ···················· | ···················· |
| L | 7971 | hCoV-19/USA/MI-QDX-510/2020\|EPI_ISL_571244\|2020-03 | ···················· | ···················· | ···················· | ···················· | ···················· | ···················· |
| L | 7972 | hCoV-19/USA/MI-QDX-511/2020\|EPI_ISL_571245\|2020-03 | ···················· | ···················· | ···················· | ···················· | ···················· | ···················· |
| L | 7973 | hCoV-19/USA/MI-QDX-512/2020\|EPI_ISL_571246\|2020-03 | ···················· | ···················· | ···················· | ···················· | ···················· | ···················· |
| L | 7974 | hCoV-19/USA/MI-QDX-515/2020\|EPI_ISL_571247\|2020-03 | ···················· | ···················· | ···················· | ···················· | ···················· | ···················· |
| L | 7975 | hCoV-19/USA/NV-QDX-418/2020\|EPI_ISL_571248\|2020-03 | ···················· | ···················· | ···················· | ···················· | ···················· | ···················· |
| L | 7976 | hCoV-19/USA/NV-QDX-419/2020\|EPI_ISL_571249\|2020-03 | ···················· | ···················· | ···················· | ···················· | ···················· | ···················· |
| L | 7977 | hCoV-19/USA/NV-QDX-420/2020\|EPI_ISL_571250\|2020-03 | ···················· | ···················· | ···················· | ···················· | ···················· | ···················· |
| L | 7978 | hCoV-19/USA/NV-QDX-421/2020\|EPI_ISL_571251\|2020-03 | ···················· | ···················· | ···················· | ···················· | ···················· | ···················· |
| L | 7979 | hCoV-19/USA/NV-QDX-432/2020\|EPI_ISL_571252\|2020-03 | ···················· | ···················· | ···················· | ···················· | ···················· | ···················· |
| L | 7980 | hCoV-19/USA/OK-QDX-430/2020\|EPI_ISL_571254\|2020-03 | ···················· | ···················· | ···················· | ···················· | ···················· | ···················· |
| L | 7981 | hCoV-19/USA/TX-QDX-413/2020\|EPI_ISL_571255\|2020-03 | ···················· | ···················· | ···················· | ···················· | ···················· | ···················· |
| L | 7982 | hCoV-19/USA/TX-QDX-414/2020\|EPI_ISL_571256\|2020-03 | ···················· | ···················· | ···················· | ···················· | ···················· | ···················· |
| L | 7983 | hCoV-19/USA/TX-QDX-415/2020\|EPI_ISL_571257\|2020-03 | ···················· | ···················· | ···················· | ···················· | ···················· | ···················· |
| L | 7984 | hCoV-19/USA/WI-QDX-438/2020\|EPI_ISL_571258\|2020-03 | ···················· | ···················· | ···················· | ···················· | ···················· | ···················· |
| L | 7985 | hCoV-19/USA/WI-QDX-439/2020\|EPI_ISL_571259\|2020-03 | ···················· | ···················· | ···················· | ···················· | ···················· | ···················· |
| L | 7986 | hCoV-19/USA/WI-QDX-440/2020\|EPI_ISL_571260\|2020-03 | ···················· | ···················· | ···················· | ···················· | ···················· | ···················· |
| L | 7987 | hCoV-19/USA/AR-QDX-532/2020\|EPI_ISL_571261\|2020-03 | ···················· | ···················· | ···················· | ···················· | ···················· | ···················· |
| L | 7988 | hCoV-19/USA/AR-QDX-537/2020\|EPI_ISL_571262\|2020-03 | ···················· | ···················· | ···················· | ···················· | ···················· | ···················· |
| L | 7989 | hCoV-19/USA/AZ-QDX-516/2020\|EPI_ISL_571263\|2020-03 | ···················· | ···················· | ···················· | ···················· | ···················· | ···················· |
| L | 7990 | hCoV-19/USA/AZ-QDX-518/2020\|EPI_ISL_571264\|2020-03 | ···················· | ···················· | ···················· | ···················· | ···················· | ···················· |
| L | 7991 | hCoV-19/USA/AZ-QDX-519/2020\|EPI_ISL_571265\|2020-03 | ···················· | ···················· | ···················· | ···················· | ···················· | ···················· |
| L | 7992 | hCoV-19/USA/AZ-QDX-520/2020\|EPI_ISL_571266\|2020-03 | ···················· | ···················· | ···················· | ···················· | ···················· | ···················· |
| L | 7993 | hCoV-19/USA/AZ-QDX-521/2020\|EPI_ISL_571267\|2020-03 | ···················· | ···················· | ···················· | ···················· | ···················· | ···················· |
| L | 7994 | hCoV-19/USA/CA-QDX-545/2020\|EPI_ISL_571268\|2020-03 | ···················· | ···················· | ···················· | ···················· | ···················· | ···················· |
| L | 7995 | hCoV-19/USA/CA-QDX-546/2020\|EPI_ISL_571269\|2020-03 | ···················· | ···················· | ···················· | ···················· | ···················· | ···················· |
| L | 7996 | hCoV-19/USA/CA-QDX-547/2020\|EPI_ISL_571270\|2020-03 | ···················· | ···················· | ···················· | ···················· | ···················· | ···················· |
| L | 7997 | hCoV-19/USA/CA-QDX-548/2020\|EPI_ISL_571271\|2020-03 | ···················· | ···················· | ···················· | ···················· | ···················· | ···················· |
| L | 7998 | hCoV-19/USA/CA-QDX-549/2020\|EPI_ISL_571272\|2020-03 | ···················· | ···················· | ···················· | ···················· | ···················· | ···················· |
| L | 7999 | hCoV-19/USA/CA-QDX-550/2020\|EPI_ISL_571273\|2020-03 | ···················· | ···················· | ···················· | ···················· | ···················· | ···················· |
| L | 8000 | hCoV-19/USA/CA-QDX-551/2020\|EPI_ISL_571274\|2020-03 | ···················· | ···················· | ···················· | ···················· | ···················· | ···················· |
| L | 8001 | hCoV-19/USA/CA-QDX-552/2020\|EPI_ISL_571275\|2020-03 | ···················· | ···················· | ···················· | ···················· | ···················· | ···················· |
| L | 8002 | hCoV-19/USA/CA-QDX-572/2020\|EPI_ISL_571280\|2020-03 | ···················· | ···················· | ···················· | ···················· | ···················· | ···················· |
| L | 8003 | hCoV-19/USA/CA-QDX-583/2020\|EPI_ISL_571281\|2020-03 | ···················· | ···················· | ···················· | ···················· | ···················· | ···················· |
| L | 8004 | hCoV-19/USA/CA-QDX-659/2020\|EPI_ISL_571283\|2020-03 | ···················· | ···················· | ···················· | ···················· | ···················· | ···················· |

| | | | | | | | | |
|---|---|---|---|---|---|---|---|---|
| L | 8005 | hCoV-19/USA/CA-QDX-661/2020\|EPI_ISL_571284\|2020-03 | .................... | .................... | .................... | .................... | .................... | .................... |
| L | 8006 | hCoV-19/USA/CA-QDX-662/2020\|EPI_ISL_571285\|2020-03 | .................... | .................... | .................... | .................... | .................... | .................... |
| L | 8007 | hCoV-19/USA/FL-QDX-574/2020\|EPI_ISL_571286\|2020-03 | .................... | .................... | .................... | .................... | .................... | .................... |
| L | 8008 | hCoV-19/USA/GA-QDX-575/2020\|EPI_ISL_571287\|2020-03 | .................... | .................... | .................... | .................... | .................... | .................... |
| L | 8009 | hCoV-19/USA/GA-QDX-577/2020\|EPI_ISL_571288\|2020-03 | .................... | .................... | .................... | .................... | .................... | .................... |
| L | 8010 | hCoV-19/USA/GA-QDX-578/2020\|EPI_ISL_571289\|2020-03 | .................... | .................... | .................... | .................... | .................... | .................... |
| L | 8011 | hCoV-19/USA/GA-QDX-579/2020\|EPI_ISL_571290\|2020-03 | .................... | .................... | .................... | .................... | .................... | .................... |
| L | 8012 | hCoV-19/USA/GA-QDX-580/2020\|EPI_ISL_571291\|2020-03 | .................... | .................... | .................... | .................... | .................... | .................... |
| L | 8013 | hCoV-19/USA/GA-QDX-581/2020\|EPI_ISL_571292\|2020-03 | .................... | .................... | .................... | .................... | .................... | .................... |
| L | 8014 | hCoV-19/USA/GA-QDX-582/2020\|EPI_ISL_571293\|2020-03 | .................... | .................... | .................... | .................... | .................... | .................... |
| L | 8015 | hCoV-19/USA/GA-QDX-594/2020\|EPI_ISL_571295\|2020-03 | .................... | .................... | .................... | .................... | .................... | .................... |
| L | 8016 | hCoV-19/USA/GA-QDX-597/2020\|EPI_ISL_571296\|2020-03 | .................... | .................... | .................... | .................... | .................... | .................... |
| L | 8017 | hCoV-19/Wales/MILK-9C73E5/2020\|EPI_ISL_573872\|2020 | .................... | .................... | .................... | .................... | .................... | .................... |
| L | 8018 | hCoV-19/Wales/PHWC-16B58B/2020\|EPI_ISL_573873\|2020 | .................... | .................... | .................... | .................... | .................... | .................... |
| L | 8019 | hCoV-19/Wales/PHWC-16B59A/2020\|EPI_ISL_573874\|2020 | .................... | .................... | .................... | .................... | .................... | .................... |
| L | 8020 | hCoV-19/Wales/PHWC-16B5B8/2020\|EPI_ISL_573875\|2020 | .................... | .................... | .................... | .................... | .................... | .................... |
| L | 8021 | hCoV-19/Wales/PHWC-16B5C7/2020\|EPI_ISL_573876\|2020 | .................... | .................... | .................... | .................... | .................... | .................... |
| L | 8022 | hCoV-19/Wales/PHWC-16B5D6/2020\|EPI_ISL_573877\|2020 | .................... | .................... | .................... | .................... | .................... | .................... |
| L | 8023 | hCoV-19/Wales/PHWC-16B5E5/2020\|EPI_ISL_573878\|2020 | .................... | .................... | .................... | .................... | .................... | .................... |
| L | 8024 | hCoV-19/Wales/PHWC-16B600/2020\|EPI_ISL_573879\|2020 | .................... | .................... | .................... | .................... | .................... | .................... |
| L | 8025 | hCoV-19/Wales/PHWC-16B61F/2020\|EPI_ISL_573880\|2020 | .................... | .................... | .................... | .................... | .................... | .................... |
| L | 8026 | hCoV-19/Wales/PHWC-16B62E/2020\|EPI_ISL_573881\|2020 | .................... | .................... | .................... | .................... | .................... | .................... |
| L | 8027 | hCoV-19/Wales/PHWC-16B63D/2020\|EPI_ISL_573882\|2020 | .................... | .................... | .................... | .................... | .................... | .................... |
| L | 8028 | hCoV-19/Wales/PHWC-16B64C/2020\|EPI_ISL_573883\|2020 | .................... | .................... | .................... | .................... | .................... | .................... |
| L | 8029 | hCoV-19/Wales/PHWC-16B679/2020\|EPI_ISL_573884\|2020 | .................... | .................... | .................... | .................... | .................... | .................... |
| L | 8030 | hCoV-19/Wales/PHWC-16B688/2020\|EPI_ISL_573885\|2020 | .................... | .................... | .................... | .................... | .................... | .................... |
| L | 8031 | hCoV-19/Wales/PHWC-16B697/2020\|EPI_ISL_573886\|2020 | .................... | .................... | .................... | .................... | .................... | .................... |
| L | 8032 | hCoV-19/Wales/PHWC-16B6B5/2020\|EPI_ISL_573887\|2020 | .................... | .................... | .................... | .................... | .................... | .................... |
| L | 8033 | hCoV-19/Wales/PHWC-16B6C4/2020\|EPI_ISL_573888\|2020 | .................... | .................... | .................... | .................... | .................... | .................... |
| L | 8034 | hCoV-19/Wales/PHWC-16B6D3/2020\|EPI_ISL_573889\|2020 | .................... | .................... | .................... | .................... | ..K................. | .................... |
| L | 8035 | hCoV-19/Wales/PHWC-16B6E2/2020\|EPI_ISL_573890\|2020 | .................... | .................... | .................... | .................... | .................... | .................... |
| L | 8036 | hCoV-19/Wales/PHWC-16B70D/2020\|EPI_ISL_573891\|2020 | .................... | .................... | .................... | .................... | .................... | .................... |
| L | 8037 | hCoV-19/Wales/PHWC-16B71C/2020\|EPI_ISL_573892\|2020 | .................... | .................... | .................... | .................... | .................... | .................... |
| L | 8038 | hCoV-19/Wales/PHWC-16B72B/2020\|EPI_ISL_573893\|2020 | .................... | .................... | .................... | .................... | .................... | .................... |
| L | 8039 | hCoV-19/Wales/PHWC-16B73A/2020\|EPI_ISL_573894\|2020 | .................... | .................... | .................... | .................... | .................... | .................... |
| L | 8040 | hCoV-19/Wales/PHWC-16B749/2020\|EPI_ISL_573895\|2020 | .................... | .................... | .................... | .................... | .................... | .................... |
| L | 8041 | hCoV-19/Wales/PHWC-16B758/2020\|EPI_ISL_573896\|2020 | .................... | .................... | .................... | .................... | .................... | .................... |
| L | 8042 | hCoV-19/Wales/PHWC-16B767/2020\|EPI_ISL_573897\|2020 | .................... | .................... | .................... | .................... | .................... | .................... |
| L | 8043 | hCoV-19/Wales/PHWC-16B776/2020\|EPI_ISL_573898\|2020 | .................... | .................... | .................... | .................... | .................... | .................... |
| L | 8044 | hCoV-19/Wales/PHWC-16B785/2020\|EPI_ISL_573899\|2020 | .................... | .................... | .................... | .................... | .................... | .................... |
| L | 8045 | hCoV-19/Wales/MILK-9E11CD/2020\|EPI_ISL_577401\|2020 | .................... | .................... | .................... | .................... | .................... | .................... |
| L | 8046 | hCoV-19/Wales/MILK-9E14D3/2020\|EPI_ISL_577402\|2020 | .................... | .................... | .................... | .................... | .................... | .................... |
| L | 8047 | hCoV-19/Wales/MILK-9E14F1/2020\|EPI_ISL_577403\|2020 | .................... | .................... | .................... | .................... | .................... | .................... |
| L | 8048 | hCoV-19/Wales/MILK-9E151C/2020\|EPI_ISL_577404\|2020 | .................... | .................... | .................... | .................... | .................... | .................... |
| L | 8049 | hCoV-19/Wales/MILK-9E152B/2020\|EPI_ISL_577405\|2020 | .................... | .................... | .................... | .................... | .................... | .................... |
| L | 8050 | hCoV-19/Wales/MILK-9E153A/2020\|EPI_ISL_577406\|2020 | .................... | .................... | .................... | .................... | .................... | .................... |
| L | 8051 | hCoV-19/Wales/MILK-9E1549/2020\|EPI_ISL_577407\|2020 | .................... | .................... | .................... | .................... | .................... | .................... |
| L | 8052 | hCoV-19/Wales/MILK-9E1558/2020\|EPI_ISL_577408\|2020 | .................... | .................... | .................... | .................... | .................... | .................... |
| L | 8053 | hCoV-19/Wales/MILK-9E17BC/2020\|EPI_ISL_577409\|2020 | .................... | .................... | .................... | .................... | .................... | .................... |
| L | 8054 | hCoV-19/Wales/MILK-9E1813/2020\|EPI_ISL_577410\|2020 | .................... | .................... | .................... | .................... | .................... | .................... |
| L | 8055 | hCoV-19/Wales/MILK-9E186E/2020\|EPI_ISL_577411\|2020 | .................... | .................... | .................... | .................... | .................... | .................... |
| L | 8056 | hCoV-19/Wales/MILK-9E189B/2020\|EPI_ISL_577413\|2020 | .................... | .................... | .................... | .................... | .................... | .................... |
| L | 8057 | hCoV-19/Wales/MILK-9E18AA/2020\|EPI_ISL_577414\|2020 | .................... | .................... | .................... | .................... | .................... | .................... |
| L | 8058 | hCoV-19/Wales/MILK-9E18B9/2020\|EPI_ISL_577415\|2020 | .................... | .................... | .................... | .................... | .................... | .................... |
| L | 8059 | hCoV-19/Wales/MILK-9E18D7/2020\|EPI_ISL_577416\|2020 | .................... | .................... | .................... | .................... | .................... | .................... |
| L | 8060 | hCoV-19/Wales/PHWC-16D5D4/2020\|EPI_ISL_577417\|2020 | .................... | .................... | .................... | .................... | .................... | .................... |
| L | 8061 | hCoV-19/Wales/PHWC-16D5E3/2020\|EPI_ISL_577418\|2020 | .................... | .................... | .................... | .................... | .................... | .................... |
| L | 8062 | hCoV-19/Wales/PHWC-16D5F2/2020\|EPI_ISL_577419\|2020 | .................... | .................... | .................... | .................... | .................... | .................... |
| L | 8063 | hCoV-19/Wales/PHWC-16D60E/2020\|EPI_ISL_577420\|2020 | .................... | .................... | .................... | .................... | .................... | .................... |
| L | 8064 | hCoV-19/Wales/PHWC-16D61D/2020\|EPI_ISL_577421\|2020 | .................... | .................... | .................... | .................... | .................... | .................... |
| L | 8065 | hCoV-19/Wales/PHWC-16D62C/2020\|EPI_ISL_577422\|2020 | .................... | .................... | .................... | .................... | .................... | .................... |
| L | 8066 | hCoV-19/Wales/PHWC-16D63B/2020\|EPI_ISL_577423\|2020 | .................... | .................... | .................... | .................... | .................... | .................... |
| L | 8067 | hCoV-19/Wales/PHWC-16D64A/2020\|EPI_ISL_577424\|2020 | .................... | .................... | .................... | .................... | .................... | .................... |
| L | 8068 | hCoV-19/Wales/PHWC-16D659/2020\|EPI_ISL_577425\|2020 | .................... | .................... | .................... | .................... | .................... | .................... |
| L | 8069 | hCoV-19/Wales/PHWC-16D668/2020\|EPI_ISL_577426\|2020 | .................... | .................... | .................... | .................... | .................... | .................... |
| L | 8070 | hCoV-19/Wales/PHWC-16D677/2020\|EPI_ISL_577427\|2020 | .................... | .................... | .................... | .................... | .................... | .................... |
| L | 8071 | hCoV-19/Wales/PHWC-16D686/2020\|EPI_ISL_577428\|2020 | .................... | .................... | .................... | .................... | .................... | .................... |
| L | 8072 | hCoV-19/Wales/PHWC-16D695/2020\|EPI_ISL_577429\|2020 | .................... | .................... | .................... | .................... | .................... | .................... |
| L | 8073 | hCoV-19/Wales/PHWC-16D6A4/2020\|EPI_ISL_577430\|2020 | .................... | .................... | .................... | .................... | .................... | .................... |
| L | 8075 | hCoV-19/Wales/PHWC-16D6C2/2020\|EPI_ISL_577432\|2020 | .................... | .................... | .................... | .................... | .................... | .................... |
| L | 8076 | hCoV-19/Wales/PHWC-16D6D1/2020\|EPI_ISL_577433\|2020 | .................... | .................... | .................... | .................... | .................... | .................... |
| L | 8077 | hCoV-19/Wales/PHWC-16D6E0/2020\|EPI_ISL_577434\|2020 | .................... | .................... | .................... | .................... | .................... | .................... |
| L | 8078 | hCoV-19/USA/LA-EVTL297/2020\|EPI_ISL_578704\|2020-05 | .................... | .................... | .................... | .................... | .................... | .................... |
| L | 8079 | hCoV-19/USA/LA-EVTL298/2020\|EPI_ISL_578705\|2020-05 | .................... | .................... | .................... | .................... | .................... | .................... |
| L | 8080 | hCoV-19/USA/LA-EVTL299/2020\|EPI_ISL_578706\|2020-05 | .................... | .................... | .................... | .................... | .................... | .................... |
| L | 8081 | hCoV-19/USA/LA-EVTL300/2020\|EPI_ISL_578707\|2020-05 | .................... | .................... | .................... | .................... | .................... | .................... |
| L | 8082 | hCoV-19/USA/LA-EVTL301/2020\|EPI_ISL_578708\|2020-05 | .................... | .................... | .................... | .................... | .................... | .................... |
| L | 8083 | hCoV-19/USA/LA-EVTL302/2020\|EPI_ISL_578709\|2020-05 | .................... | .................... | .................... | .................... | .................... | .................... |
| L | 8084 | hCoV-19/USA/LA-EVTL303/2020\|EPI_ISL_578710\|2020-05 | .................... | .................... | .................... | .................... | .................... | .................... |
| L | 8085 | hCoV-19/USA/LA-EVTL304/2020\|EPI_ISL_578711\|2020-05 | .................... | .................... | .................... | .................... | .................... | .................... |

| | | | | | | | | |
|---|---|---|---|---|---|---|---|---|
| L | 8086 | hCoV-19/USA/LA-EVTL308/2020\|EPI_ISL_578715\|2020-05 | ···················· | ···················· | ····················· | ···················· | ···················· | ···················· |
| L | 8087 | hCoV-19/USA/LA-EVTL316/2020\|EPI_ISL_578723\|2020-05 | ···················· | ···················· | ····················· | ···················· | ···················· | ···················· |
| L | 8088 | hCoV-19/USA/LA-EVTL317/2020\|EPI_ISL_578724\|2020-05 | ···················· | ···················· | ····················· | ···················· | ···················· | ···················· |
| L | 8089 | hCoV-19/USA/LA-EVTL318/2020\|EPI_ISL_578725\|2020-05 | ···················· | ···················· | ····················· | ···················· | ···················· | ···················· |
| L | 8090 | hCoV-19/USA/LA-EVTL319/2020\|EPI_ISL_578726\|2020-05 | ···················· | ···················· | ····················· | ···················· | ···················· | ···················· |
| L | 8091 | hCoV-19/USA/LA-EVTL320/2020\|EPI_ISL_578727\|2020-05 | ···················· | ···················· | ····················· | ···················· | ···················· | ···················· |
| L | 8092 | hCoV-19/USA/LA-EVTL322/2020\|EPI_ISL_578728\|2020-05 | ···················· | ···················· | ····················· | ···················· | ···················· | ···················· |
| L | 8093 | hCoV-19/USA/LA-EVTL323/2020\|EPI_ISL_578729\|2020-05 | ···················· | ···················· | ····················· | ···················· | ···················· | ···················· |
| L | 8094 | hCoV-19/USA/LA-EVTL324/2020\|EPI_ISL_578730\|2020-05 | ···················· | ···················· | ····················· | ···················· | ···················· | ···················· |
| L | 8095 | hCoV-19/USA/LA-EVTL325/2020\|EPI_ISL_578731\|2020-05 | ···················· | ···················· | ····················· | ···················· | ···················· | ···················· |
| L | 8096 | hCoV-19/USA/LA-EVTL326/2020\|EPI_ISL_578732\|2020-05 | ···················· | ···················· | ····················· | ···················· | ···················· | ···················· |
| L | 8097 | hCoV-19/USA/LA-EVTL327/2020\|EPI_ISL_578733\|2020-05 | ···················· | ···················· | ····················· | ···················· | ···················· | ···················· |
| L | 8098 | hCoV-19/USA/LA-EVTL328/2020\|EPI_ISL_578734\|2020-05 | ···················· | ···················· | ····················· | ···················· | ···················· | ···················· |
| L | 8099 | hCoV-19/USA/LA-EVTL329/2020\|EPI_ISL_578735\|2020-05 | ···················· | ···················· | ····················· | ···················· | ···················· | ···················· |
| L | 8100 | hCoV-19/USA/LA-EVTL330/2020\|EPI_ISL_578736\|2020-05 | ···················· | ···················· | ····················· | ···················· | ···················· | ···················· |
| L | 8101 | hCoV-19/USA/LA-EVTL331/2020\|EPI_ISL_578737\|2020-05 | ···················· | ···················· | ····················· | ···················· | ···················· | ···················· |
| L | 8102 | hCoV-19/USA/LA-EVTL332/2020\|EPI_ISL_578738\|2020-05 | ···················· | ···················· | ····················· | ···················· | ···················· | ···················· |
| L | 8103 | hCoV-19/USA/LA-EVTL333/2020\|EPI_ISL_578739\|2020-05 | ···················· | ···················· | ····················· | ···················· | ···················· | ···················· |
| L | 8104 | hCoV-19/USA/LA-EVTL334/2020\|EPI_ISL_578740\|2020-05 | ···················· | ···················· | ····················· | ···················· | ···················· | ···················· |
| L | 8105 | hCoV-19/USA/LA-EVTL335/2020\|EPI_ISL_578741\|2020-05 | ···················· | ···················· | ····················· | ···················· | ···················· | ···················· |
| L | 8106 | hCoV-19/USA/LA-EVTL336/2020\|EPI_ISL_578742\|2020-05 | ···················· | ···················· | ····················· | ···················· | ···················· | ···················· |
| L | 8107 | hCoV-19/USA/LA-EVTL337/2020\|EPI_ISL_578743\|2020-05 | ···················· | ···················· | ····················· | ···················· | ···················· | ···················· |
| L | 8108 | hCoV-19/USA/LA-EVTL338/2020\|EPI_ISL_578744\|2020-05 | ···················· | ···················· | ····················· | ···················· | ···················· | ···················· |
| L | 8109 | hCoV-19/USA/LA-EVTL339/2020\|EPI_ISL_578745\|2020-05 | ···················· | ···················· | ····················· | ···················· | ···················· | ···················· |
| L | 8110 | hCoV-19/USA/LA-EVTL340/2020\|EPI_ISL_578746\|2020-05 | ···················· | ···················· | ····················· | ···················· | ···················· | ···················· |
| L | 8111 | hCoV-19/USA/LA-EVTL341/2020\|EPI_ISL_578747\|2020-05 | ···················· | ···················· | ····················· | ···················· | ···················· | ···················· |
| L | 8112 | hCoV-19/USA/LA-EVTL342/2020\|EPI_ISL_578748\|2020-05 | ···················· | ···················· | ····················· | ···················· | ···················· | ···················· |
| L | 8113 | hCoV-19/USA/LA-EVTL343/2020\|EPI_ISL_578749\|2020-05 | ···················· | ···················· | ····················· | ···················· | ···················· | ···················· |
| L | 8114 | hCoV-19/USA/LA-EVTL344/2020\|EPI_ISL_578750\|2020-05 | ···················· | ···················· | ····················· | ···················· | ···················· | ···················· |
| L | 8115 | hCoV-19/USA/LA-EVTL345/2020\|EPI_ISL_578751\|2020-05 | ···················· | ···················· | ····················· | ···················· | ···················· | ···················· |
| L | 8116 | hCoV-19/USA/LA-EVTL346/2020\|EPI_ISL_578752\|2020-05 | ···················· | ···················· | ····················· | ···················· | ···················· | ···················· |
| L | 8117 | hCoV-19/USA/LA-EVTL347/2020\|EPI_ISL_578753\|2020-05 | ···················· | ···················· | ····················· | ···················· | ···················· | ···················· |
| L | 8118 | hCoV-19/USA/LA-EVTL348/2020\|EPI_ISL_578754\|2020-05 | ···················· | ···················· | ····················· | ···················· | ···················· | ···················· |
| L | 8119 | hCoV-19/USA/LA-EVTL349/2020\|EPI_ISL_578755\|2020-05 | ···················· | ···················· | ····················· | ···················· | ···················· | ···················· |
| L | 8120 | hCoV-19/USA/LA-EVTL350/2020\|EPI_ISL_578756\|2020-05 | ···················· | ···················· | ····················· | ···················· | ···················· | ···················· |
| L | 8121 | hCoV-19/USA/LA-EVTL351/2020\|EPI_ISL_578757\|2020-05 | ···················· | ···················· | ····················· | ···················· | ···················· | ···················· |
| L | 8122 | hCoV-19/USA/LA-EVTL352/2020\|EPI_ISL_578758\|2020-05 | ···················· | ···················· | ····················· | ···················· | ···················· | ···················· |
| L | 8123 | hCoV-19/USA/LA-EVTL353/2020\|EPI_ISL_578759\|2020-05 | ···················· | ···················· | ····················· | ···················· | ···················· | ···················· |
| L | 8124 | hCoV-19/USA/LA-EVTL354/2020\|EPI_ISL_578760\|2020-05 | ···················· | ···················· | ····················· | ···················· | ···················· | ···················· |
| L | 8125 | hCoV-19/USA/LA-EVTL355/2020\|EPI_ISL_578761\|2020-05 | ···················· | ···················· | ····················· | ···················· | ···················· | ···················· |
| L | 8126 | hCoV-19/USA/LA-EVTL356/2020\|EPI_ISL_578762\|2020-05 | ···················· | ···················· | ····················· | ···················· | ···················· | ···················· |
| L | 8127 | hCoV-19/USA/LA-EVTL357/2020\|EPI_ISL_578763\|2020-05 | ···················· | ···················· | ····················· | ···················· | ···················· | ···················· |
| L | 8128 | hCoV-19/USA/LA-EVTL358/2020\|EPI_ISL_578764\|2020-05 | ···················· | ···················· | ····················· | ···················· | ···················· | ···················· |
| L | 8129 | hCoV-19/USA/LA-EVTL359/2020\|EPI_ISL_578765\|2020-05 | ···················· | ···················· | ····················· | ···················· | ···················· | ···················· |
| L | 8130 | hCoV-19/USA/LA-EVTL360/2020\|EPI_ISL_578766\|2020-05 | ···················· | ···················· | ····················· | ···················· | ···················· | ···················· |
| L | 8131 | hCoV-19/USA/CA-CZB-192/2020\|EPI_ISL_583200\|2020-08 | ···················· | ···················· | ····················· | ···················· | ···················· | ···················· |
| L | 8132 | hCoV-19/USA/CA-CZB-191/2020\|EPI_ISL_583201\|2020-08 | ···················· | ···················· | ····················· | ···················· | ···················· | ···················· |
| L | 8133 | hCoV-19/USA/CA-CZB-169/2020\|EPI_ISL_583202\|2020-08 | ···················· | ·A·················· | ····················· | ···················· | ···················· | ···················· |
| L | 8134 | hCoV-19/USA/CA-CZB-168/2020\|EPI_ISL_583203\|2020-08 | ···················· | ·A·················· | ····················· | ···················· | ···················· | ···················· |
| L | 8135 | hCoV-19/USA/CA-CZB-165/2020\|EPI_ISL_583204\|2020-09 | ···················· | ···················· | ····················· | ···················· | ···················· | ···················· |
| L | 8136 | hCoV-19/USA/CA-CZB-164/2020\|EPI_ISL_583205\|2020-09 | ···················· | ···················· | ····················· | ···················· | ···················· | ···················· |
| L | 8137 | hCoV-19/USA/CA-CZB-163/2020\|EPI_ISL_583206\|2020-09 | ···················· | ···················· | ····················· | ···················· | ···················· | ···················· |
| L | 8138 | hCoV-19/USA/CA-CZB-162/2020\|EPI_ISL_583207\|2020-09 | ···················· | ···················· | ····················· | ···················· | ···················· | ···················· |
| L | 8139 | hCoV-19/USA/CA-CZB-160/2020\|EPI_ISL_583208\|2020-08 | ···················· | ···················· | ····················· | ···················· | ···················· | ···················· |
| L | 8140 | hCoV-19/USA/CA-CZB-159/2020\|EPI_ISL_583209\|2020-08 | ···················· | ···················· | ····················· | ···················· | ···················· | ···················· |
| L | 8141 | hCoV-19/USA/CA-CZB-5851/2020\|EPI_ISL_583210\|2020-0 | ···················· | ···················· | ····················· | ···················· | ···················· | ···················· |
| L | 8142 | hCoV-19/USA/CA-CZB-5842/2020\|EPI_ISL_583211\|2020-0 | ···················· | ···················· | ····················· | ···················· | ···················· | ···················· |
| L | 8143 | hCoV-19/USA/CA-CZB-5839/2020\|EPI_ISL_583212\|2020-0 | ···················· | ···················· | ····················· | ···················· | ···················· | ···················· |
| L | 8144 | hCoV-19/USA/CA-CZB-5695/2020\|EPI_ISL_583213\|2020-0 | ···················· | ·A·················· | ····················· | ···················· | ···················· | ···················· |
| L | 8145 | hCoV-19/USA/CA-CZB-5693/2020\|EPI_ISL_583214\|2020-0 | ···················· | ···················· | ····················· | ···················· | ·············T······ | ···················· |
| L | 8146 | hCoV-19/USA/CA-CZB-5691/2020\|EPI_ISL_583215\|2020-0 | ···················· | ···················· | ····················· | ···················· | ···················· | ···················· |
| L | 8147 | hCoV-19/USA/CA-CZB-5670/2020\|EPI_ISL_583216\|2020-0 | ···················· | ···················· | ····················· | ···················· | ···················· | ···················· |
| L | 8148 | hCoV-19/USA/CA-CZB-5668/2020\|EPI_ISL_583217\|2020-0 | ···················· | ···················· | ····················· | ···················· | ···················· | ···················· |
| L | 8149 | hCoV-19/USA/CA-CZB-5667/2020\|EPI_ISL_583218\|2020-0 | ···················· | ·A·················· | ····················· | ···················· | ···················· | ···················· |
| L | 8150 | hCoV-19/USA/CA-CZB-5174/2020\|EPI_ISL_583219\|2020-0 | ···················· | ·A·················· | ····················· | ···················· | ···················· | ···················· |
| L | 8151 | hCoV-19/USA/CA-CZB-5171/2020\|EPI_ISL_583220\|2020-0 | ···················· | ·A·················· | ····················· | ···················· | ···················· | ···················· |
| L | 8152 | hCoV-19/USA/CA-CZB-5170/2020\|EPI_ISL_583221\|2020-0 | ···················· | ···················· | ··T·················· | ···················· | ···················· | ···················· |
| L | 8153 | hCoV-19/USA/CA-CZB-5168/2020\|EPI_ISL_583222\|2020-0 | ···················· | ·A·················· | ····················· | ···················· | ···················· | ···················· |
| L | 8154 | hCoV-19/USA/CA-CZB-5167/2020\|EPI_ISL_583223\|2020-0 | ···················· | ···················· | ····················· | ···················· | ···················· | ···················· |
| L | 8155 | hCoV-19/USA/CA-CZB-5159/2020\|EPI_ISL_583224\|2020-0 | ···················· | ···················· | ····················· | ···················· | ···················· | ···················· |
| L | 8156 | hCoV-19/USA/CA-CZB-7161/2020\|EPI_ISL_583225\|2020-0 | ···················· | ·A·················· | ····················· | ···················· | ···················· | ···················· |
| L | 8157 | hCoV-19/USA/CA-CZB-7160/2020\|EPI_ISL_583226\|2020-0 | ···················· | ···················· | ····················· | ···················· | ···················· | ···················· |
| L | 8158 | hCoV-19/USA/CA-CZB-7158/2020\|EPI_ISL_583227\|2020-0 | ···················· | ···················· | ····················· | ···················· | ···················· | ···················· |
| L | 8159 | hCoV-19/USA/CA-CZB-7157/2020\|EPI_ISL_583228\|2020-0 | ···················· | ···················· | ····················· | ···················· | ···················· | ···················· |
| L | 8160 | hCoV-19/USA/DE-DHSS-F917889/2020\|EPI_ISL_583246\|20 | ···················· | ···················· | ····················· | ···················· | ···················· | ···················· |
| L | 8161 | hCoV-19/USA/DE-DHSS-F917866/2020\|EPI_ISL_583248\|20 | ···················· | ···················· | ····················· | ···················· | ···················· | ···················· |
| L | 8167 | hCoV-19/USA/MI-UM-10035622031/2020\|EPI_ISL_583260\| | ···················· | ···················· | ····················· | ···················· | ···················· | ···················· |
| L | 8170 | hCoV-19/USA/MI-UM-10035702532/2020\|EPI_ISL_583263\| | ···················· | ·A·················· | ····················· | ···················· | ···················· | ···················· |
| L | 8171 | hCoV-19/USA/MI-UM-10035555295/2020\|EPI_ISL_583264\| | ···················· | ·A·················· | ····················· | ···················· | ···················· | ···················· |
| L | 8172 | hCoV-19/USA/MI-UM-10035692136/2020\|EPI_ISL_583266\| | ···················· | ·A·················· | ····················· | ···················· | ···················· | ···················· |

```
L 8175 hCoV-19/USA/MI-UM-10035702585/2020|EPI_ISL_583269| ···················· ···················· ···················· ···················· ···················· ····················
L 8176 hCoV-19/USA/MI-UM-10035615810/2020|EPI_ISL_583270| ···················· ···················· ···················· ···················· ···················· ····················
L 8177 hCoV-19/USA/MI-UM-10035638276/2020|EPI_ISL_583271| ···················· ·············A······ ···················· ···················· ···················· ····················
L 8180 hCoV-19/USA/MI-UM-10035681864/2020|EPI_ISL_583274| ···················· ·A·················· ···················· ···················· ···················· ····················
L 8181 hCoV-19/USA/MI-UM-10035531897/2020|EPI_ISL_583275| ···················· ···················· ···················· ···················· ···················· ····················
L 8182 hCoV-19/USA/MI-UM-10035711552/2020|EPI_ISL_583276| ···················· ···················· ···················· ···················· ···················· ····················
L 8183 hCoV-19/USA/MI-UM-10035701585/2020|EPI_ISL_583288| ···················· ···················· ···················· ···················· ···················· ····················
L 8184 hCoV-19/USA/MI-UM-10035739693/2020|EPI_ISL_583289| ···················· ···················· ···················· ···················· ···················· ····················
L 8187 hCoV-19/USA/MI-UM-10035745470/2020|EPI_ISL_583293| ···················· ·A·················· ···················· ···················· ···················· ····················
L 8188 hCoV-19/USA/MI-UM-10035738235/2020|EPI_ISL_583294| ···················· ·············A······ ···················· ···················· ···················· ····················
L 8189 hCoV-19/USA/MI-UM-10035732796/2020|EPI_ISL_583295| T··················· ·A·················· ···················· ···················· ···················· ····················
L 8190 hCoV-19/USA/MI-UM-10035726990/2020|EPI_ISL_583296| ···················· ·A·················· ···················· ···················· ···················· ····················
L 8191 hCoV-19/USA/MI-UM-10035741674/2020|EPI_ISL_583297| ···················· ···················· ···················· ···················· ···················· ····················
L 8193 hCoV-19/USA/CA-QDX-553/2020|EPI_ISL_571276|2020-03 ···················· ···················· ···················· ···················· ···················· ····················
L 8194 hCoV-19/USA/CA-QDX-554/2020|EPI_ISL_571277|2020-03 ···················· ···················· ···················· ···················· ···················· ····················
L 8195 hCoV-19/USA/CA-QDX-556/2020|EPI_ISL_571278|2020-03 ···················· ···················· ···················· ···················· ···················· ····················
L 8196 hCoV-19/USA/CA-QDX-557/2020|EPI_ISL_571279|2020-03 ···················· ···················· ···················· ···················· ···················· ····················
L 8197 hCoV-19/England/SHEF-C4E68/2020|EPI_ISL_573700|202 ···················· ···················· ···················· ···················· ···················· ····················
L 8198 hCoV-19/England/SHEF-C531D/2020|EPI_ISL_573701|202 ···················· ···················· ···················· ···················· ···················· ····················
L 8199 hCoV-19/England/SHEF-C5562/2020|EPI_ISL_573702|202 ···················· ···················· ···················· ···················· ···················· ····················
L 8200 hCoV-19/England/SHEF-C55BD/2020|EPI_ISL_573703|202 ···················· ···················· ···················· ···················· ···················· ····················
L 8201 hCoV-19/England/SHEF-C56F6/2020|EPI_ISL_573704|202 ···················· ·A·······A·········· ···················· ···················· ···················· ····················
L 8202 hCoV-19/England/SHEF-C5CC7/2020|EPI_ISL_573705|202 ···················· ···················· ···················· ···················· ···················· ····················
L 8203 hCoV-19/England/SHEF-C5CE5/2020|EPI_ISL_573706|202 ···················· ···················· ···················· ···················· ···················· ····················
L 8204 hCoV-19/England/SHEF-C5E49/2020|EPI_ISL_573707|202 ···················· ···················· ···················· ···················· ···················· ····················
L 8205 hCoV-19/England/SHEF-C6D5A/2020|EPI_ISL_573708|202 ···················· ···················· ···················· ···················· ···················· ····················
L 8206 hCoV-19/England/SHEF-C6F63/2020|EPI_ISL_573709|202 ···················· ···················· ···················· ···················· ···················· ····················
L 8207 hCoV-19/England/SHEF-C7445/2020|EPI_ISL_573710|202 ···················· ···················· ···················· ···················· ···················· ····················
L 8208 hCoV-19/England/SHEF-C7490/2020|EPI_ISL_573711|202 ···················· ···················· ···················· ···················· ···················· ····················
L 8209 hCoV-19/England/SHEF-C773D/2020|EPI_ISL_573712|202 ···················· ···················· ···················· ···················· ···················· ····················
L 8210 hCoV-19/England/SHEF-C775B/2020|EPI_ISL_573713|202 ···················· ···················· ···················· ···················· ···················· ····················
L 8211 hCoV-19/England/SHEF-C7DC2/2020|EPI_ISL_573714|202 ···················· ···················· ···················· ···················· ···················· ····················
L 8212 hCoV-19/England/SHEF-C8972/2020|EPI_ISL_573715|202 ···················· ···················· ···················· ···················· ···················· ····················
L 8213 hCoV-19/England/SHEF-C8A24/2020|EPI_ISL_573716|202 ···················· ···················· ···················· ···················· ···················· ····················
L 8214 hCoV-19/England/SHEF-C8DA3/2020|EPI_ISL_573717|202 ···················· ···················· ···················· ···················· ···················· ····················
L 8215 hCoV-19/England/SHEF-C90B8/2020|EPI_ISL_573718|202 ···················· ···················· ···················· ···················· ···················· ····················
L 8216 hCoV-19/England/SHEF-C94CB/2020|EPI_ISL_573719|202 ···················· ···················· ···················· ···················· ···················· ····················
L 8217 hCoV-19/England/SHEF-C95E6/2020|EPI_ISL_573720|202 ···················· ···················· ···················· ···················· ···················· ····················
L 8218 hCoV-19/England/SHEF-C97C2/2020|EPI_ISL_573721|202 ···················· ···················· ···················· ···················· ···················· ····················
L 8219 hCoV-19/England/SHEF-CA424/2020|EPI_ISL_573722|202 ···················· ···················· ···················· ···················· ···················· ····················
L 8220 hCoV-19/England/SHEF-CAB10/2020|EPI_ISL_573723|202 ···················· ···················· ···················· ···················· ···················· ····················
L 8221 hCoV-19/England/SHEF-CAD65/2020|EPI_ISL_573724|202 ···················· ···················· ···················· ···················· ···················· ····················
L 8222 hCoV-19/England/SHEF-CAEEA/2020|EPI_ISL_573725|202 ···················· ·A·················· ···················· ···················· ···················· ····················
L 8223 hCoV-19/England/SHEF-CB274/2020|EPI_ISL_573726|202 ···················· ···················· ···················· ···················· ···················· ····················
L 8224 hCoV-19/England/SHEF-CB8AF/2020|EPI_ISL_573727|202 ···················· ···················· ···················· ···················· ···················· ····················
L 8225 hCoV-19/England/SHEF-CBAA9/2020|EPI_ISL_573728|202 ···················· ···················· ···················· ···················· ···················· ····················
L 8226 hCoV-19/England/SHEF-CBC85/2020|EPI_ISL_573729|202 ···················· ···················· ···················· ···················· ···················· ····················
L 8227 hCoV-19/England/SHEF-CBD55/2020|EPI_ISL_573730|202 ···················· ···················· ···················· ···················· ······T············· ····················
L 8228 hCoV-19/England/SHEF-CBE61/2020|EPI_ISL_573731|202 ···················· ···················· ···················· ···················· ···················· ····················
L 8229 hCoV-19/England/SHEF-CC835/2020|EPI_ISL_573732|202 ···················· ···················· ···················· ···················· ···················· ····················
L 8230 hCoV-19/England/SHEF-CCF03/2020|EPI_ISL_573733|202 ···················· ······C············· ···················· ···················· ···················· ····················
L 8231 hCoV-19/England/SHEF-CD2EB/2020|EPI_ISL_573734|202 ···················· ···················· ···················· ···················· ···················· ····················
L 8232 hCoV-19/England/SHEF-CD360/2020|EPI_ISL_573735|202 ···················· ···················· ···················· ···················· ···················· ····················
L 8233 hCoV-19/England/SHEF-CDAB6/2020|EPI_ISL_573736|202 ···················· ···················· ···················· ···················· ···················· ····················
L 8234 hCoV-19/England/SHEF-CE745/2020|EPI_ISL_573737|202 ···················· ···················· ···················· ···················· ···················· ····················
L 8235 hCoV-19/England/SHEF-CE87F/2020|EPI_ISL_573738|202 ···················· ···················· ···················· ···················· ···················· ····················
L 8236 hCoV-19/England/SHEF-CE9F4/2020|EPI_ISL_573739|202 ···················· ···················· ···················· ···················· ···················· ····················
L 8237 hCoV-19/England/SHEF-CEB76/2020|EPI_ISL_573740|202 ···················· ···················· ···················· ···················· ···················· ····················
L 8238 hCoV-19/England/SHEF-CEE6E/2020|EPI_ISL_573741|202 ···················· ···················· ···················· ···················· ···················· ····················
L 8239 hCoV-19/England/SHEF-CEE9B/2020|EPI_ISL_573742|202 ···················· ···················· ···················· ···················· ···················· ····················
L 8240 hCoV-19/England/SHEF-CF0A3/2020|EPI_ISL_573743|202 ···················· ···················· ···················· ···················· ···················· ····················
L 8241 hCoV-19/England/SHEF-CF1FB/2020|EPI_ISL_573744|202 ···················· ···················· ···················· ···················· ···················· ····················
L 8242 hCoV-19/England/SHEF-CF3AA/2020|EPI_ISL_573745|202 ···················· ···················· ···················· ···················· ···················· ····················
L 8243 hCoV-19/England/SHEF-CF4C5/2020|EPI_ISL_573746|202 ···················· ···················· ···················· ···················· ···················· ····················
L 8244 hCoV-19/England/SHEF-CF692/2020|EPI_ISL_573747|202 ···················· ···················· ···················· ···················· ···················· ····················
L 8245 hCoV-19/England/SHEF-CF920/2020|EPI_ISL_573748|202 ···················· ···················· ···················· ···················· ···················· ····················
L 8246 hCoV-19/England/SHEF-D0B9F/2020|EPI_ISL_573749|202 ···················· ···················· ···················· ···················· ···················· ····················
L 8247 hCoV-19/England/SHEF-D12E4/2020|EPI_ISL_573750|202 ···················· ···················· ···················· ···················· ···················· ····················
L 8248 hCoV-19/England/QEUH-9AE4E1/2020|EPI_ISL_577221|20 ···················· ···················· ···················· ···················· ···················· ····················
L 8249 hCoV-19/England/QEUH-9AE51B/2020|EPI_ISL_577224|20 ···················· ···················· ···················· ···················· ···················· ····················
L 8250 hCoV-19/England/QEUH-9AE539/2020|EPI_ISL_577225|20 ···················· ···················· ···················· ···················· ···················· ····················
L 8251 hCoV-19/England/QEUH-9AE5FD/2020|EPI_ISL_577227|20 ···················· ···················· ···················· ···················· ···················· ····················
L 8252 hCoV-19/England/QEUH-9AE645/2020|EPI_ISL_577230|20 ···················· ···················· ···················· ···················· ···················· ····················
L 8253 hCoV-19/England/QEUH-9AE663/2020|EPI_ISL_577231|20 ···················· ···················· ···················· ···················· ···················· ····················
L 8254 hCoV-19/England/QEUH-9AE6EB/2020|EPI_ISL_577234|20 ···················· ···················· ···················· ···················· ···················· ····················
L 8256 hCoV-19/England/QEUH-9AE742/2020|EPI_ISL_577236|20 ···················· ···················· ···················· ···················· ···················· ····················
L 8257 hCoV-19/England/QEUH-9AE79D/2020|EPI_ISL_577238|20 ···················· ···················· ···················· ···················· ···················· ····················
L 8258 hCoV-19/England/QEUH-9AE7AC/2020|EPI_ISL_577239|20 ···················· ···················· ···················· ···················· ···················· ····················
L 8259 hCoV-19/England/QEUH-9AE7E8/2020|EPI_ISL_577242|20 ···················· ···················· ···················· ···················· ···················· ····················
L 8261 hCoV-19/England/QEUH-9AE8B8/2020|EPI_ISL_577251|20 ···················· ···················· ···················· ···················· ···················· ····················
```

| | | | | | | | | |
|---|---|---|---|---|---|---|---|---|
| L | 8262 | hCoV-19/England/QEUH-9AE8C7/2020\|EPI_ISL_577252\|20 | ···················· | ···················· | ····················· | ···················· | ···················· | ···················· |
| L | 8263 | hCoV-19/England/QEUH-9AE92E/2020\|EPI_ISL_577255\|20 | ···················· | ···················· | ····················· | ···················· | ···················· | ···················· |
| L | 8264 | hCoV-19/England/QEUH-9AE997/2020\|EPI_ISL_577258\|20 | ···················· | ···················· | ····················· | ···················· | ···················· | ···················· |
| L | 8266 | hCoV-19/England/QEUH-9AEA67/2020\|EPI_ISL_577270\|20 | ···················· | ·A·················· | ····················· | ···················· | ···················· | ···················· |
| L | 8268 | hCoV-19/England/QEUH-9AEAA3/2020\|EPI_ISL_577272\|20 | ···················· | ···················· | ····················· | ···················· | ···················· | ···················· |
| L | 8269 | hCoV-19/England/QEUH-9D1069/2020\|EPI_ISL_577273\|20 | ···················· | ···················· | ····················· | ···················· | ···················· | ···················· |
| L | 8270 | hCoV-19/England/QEUH-9D1087/2020\|EPI_ISL_577274\|20 | ···················· | ···················· | ····················· | ···················· | ···················· | ···················· |
| L | 8271 | hCoV-19/England/QEUH-9D1227/2020\|EPI_ISL_577276\|20 | ···················· | ···················· | ····················· | ···················· | ···················· | ···················· |
| L | 8272 | hCoV-19/England/QEUH-9D1290/2020\|EPI_ISL_577279\|20 | ···················· | ·A·················· | ····················· | ···················· | ···················· | ···················· |
| L | 8273 | hCoV-19/England/QEUH-9D1351/2020\|EPI_ISL_577280\|20 | ···················· | ···················· | ····················· | ···················· | ···················· | ···················· |
| L | 8274 | hCoV-19/England/QEUH-9D1360/2020\|EPI_ISL_577281\|20 | ···················· | ···················· | ····················· | ···················· | ···················· | ···················· |
| L | 8275 | hCoV-19/England/QEUH-9D1412/2020\|EPI_ISL_577283\|20 | ···················· | ···················· | ····················· | ···················· | ········A··········· | ···················· |
| L | 8276 | hCoV-19/England/QEUH-9D146D/2020\|EPI_ISL_577285\|20 | ···················· | ·A·················· | ····················· | ···················· | ···················· | ···················· |
| L | 8277 | hCoV-19/England/QEUH-9D149A/2020\|EPI_ISL_577286\|20 | ···················· | ···················· | ····················· | ···················· | ···················· | ···················· |
| L | 8278 | hCoV-19/England/QEUH-9D14B8/2020\|EPI_ISL_577287\|20 | ···················· | ···················· | ····················· | ···················· | ···················· | ···················· |
| L | 8279 | hCoV-19/England/QEUH-9D1500/2020\|EPI_ISL_577288\|20 | ···················· | ···················· | ····················· | ···················· | ···················· | ···················· |
| L | 8280 | hCoV-19/England/QEUH-9D152E/2020\|EPI_ISL_577289\|20 | ···················· | ···················· | ····················· | ···················· | ···················· | ···················· |
| L | 8282 | hCoV-19/England/QEUH-9D1649/2020\|EPI_ISL_577293\|20 | ···················· | ···················· | ····················· | ···················· | ···················· | ···················· |
| L | 8283 | hCoV-19/England/QEUH-9D1676/2020\|EPI_ISL_577294\|20 | ···················· | ···················· | ····················· | ···················· | ···················· | ···················· |
| L | 8284 | hCoV-19/England/QEUH-9D1685/2020\|EPI_ISL_577295\|20 | ···················· | ·A·················· | ····················· | ···················· | ···················· | ··A················· |
| L | 8285 | hCoV-19/England/QEUH-9D18F8/2020\|EPI_ISL_577296\|20 | ···················· | ···················· | ····················· | ···················· | ···················· | ···················· |
| L | 8286 | hCoV-19/England/QEUH-9D1913/2020\|EPI_ISL_577297\|20 | ···················· | ·A·················· | ····················· | ···················· | ···················· | ···················· |
| L | 8287 | hCoV-19/England/QEUH-9D1A5C/2020\|EPI_ISL_577298\|20 | ···················· | ···················· | ····················· | ···················· | ···················· | ···················· |
| L | 8288 | hCoV-19/England/QEUH-9D2CCE/2020\|EPI_ISL_577299\|20 | ···················· | ···················· | ····················· | ···················· | ···················· | ···················· |
| L | 8289 | hCoV-19/USA/WI-WSLH-200355/2020\|EPI_ISL_578525\|202 | ···················· | ···················· | ····················· | ···················· | ···················· | ···················· |
| L | 8290 | hCoV-19/USA/WI-WSLH-200356/2020\|EPI_ISL_578526\|202 | ···················· | ···················· | ····················· | ···················· | ···················· | ···················· |
| L | 8291 | hCoV-19/USA/WI-WSLH-200357/2020\|EPI_ISL_578527\|202 | ···················· | ···················· | ····················· | ···················· | ···················· | ···················· |
| L | 8292 | hCoV-19/USA/WI-WSLH-200358/2020\|EPI_ISL_578528\|202 | ···················· | ···················· | ····················· | ···················· | ···················· | ···················· |
| L | 8293 | hCoV-19/USA/WI-WSLH-200359/2020\|EPI_ISL_578529\|202 | ···················· | ···················· | ····················· | ···················· | ···················· | ···················· |
| L | 8294 | hCoV-19/USA/WI-WSLH-200360/2020\|EPI_ISL_578530\|202 | ···················· | ···················· | ····················· | ···················· | ···················· | ···················· |
| L | 8295 | hCoV-19/USA/WI-WSLH-200361/2020\|EPI_ISL_578531\|202 | ···················· | ···················· | ····················· | ···················· | ···················· | ···················· |
| L | 8296 | hCoV-19/USA/WI-WSLH-200362/2020\|EPI_ISL_578532\|202 | ···················· | ···················· | ····················· | ···················· | ···················· | ···················· |
| L | 8297 | hCoV-19/USA/WI-WSLH-200363/2020\|EPI_ISL_578533\|202 | ···················· | ···················· | ····················· | ···················· | ···················· | ···················· |
| L | 8298 | hCoV-19/USA/WI-WSLH-200364/2020\|EPI_ISL_578534\|202 | ···················· | ···················· | ····················· | ···················· | ···················· | ···················· |
| L | 8299 | hCoV-19/USA/WI-WSLH-200366/2020\|EPI_ISL_578535\|202 | ···················· | ···················· | ····················· | ···················· | ···················· | ···················· |
| L | 8301 | hCoV-19/USA/WI-WSLH-200368/2020\|EPI_ISL_578537\|202 | ···················· | ···················· | ····················· | ···················· | ···················· | ···················· |
| L | 8302 | hCoV-19/USA/WI-WSLH-200369/2020\|EPI_ISL_578538\|202 | ···················· | ···················· | ····················· | ···················· | ···················· | ···················· |
| L | 8303 | hCoV-19/USA/WI-WSLH-200370/2020\|EPI_ISL_578539\|202 | ···················· | ···················· | ····················· | ···················· | ···················· | ···················· |
| L | 8304 | hCoV-19/USA/WI-WSLH-200371/2020\|EPI_ISL_578540\|202 | ···················· | ···················· | ····················· | ···················· | ···················· | ···················· |
| L | 8305 | hCoV-19/USA/WI-WSLH-200372/2020\|EPI_ISL_578541\|202 | ···················· | ···················· | ····················· | ···················· | ···················· | ···················· |
| L | 8306 | hCoV-19/USA/WI-WSLH-200373/2020\|EPI_ISL_578542\|202 | ···················· | ···················· | ····················· | ···················· | ···················· | ···················· |
| L | 8307 | hCoV-19/USA/WI-WSLH-200374/2020\|EPI_ISL_578543\|202 | ···················· | ···················· | ····················· | ···················· | ···················· | ···················· |
| L | 8308 | hCoV-19/USA/WI-WSLH-200375/2020\|EPI_ISL_578544\|202 | ···················· | ···················· | ····················· | ···················· | ···················· | ···················· |
| L | 8310 | hCoV-19/USA/WI-WSLH-200377/2020\|EPI_ISL_578546\|202 | ···················· | ···················· | ····················· | ···················· | ···················· | ···················· |
| L | 8353 | hCoV-19/USA/WI-WSLH-200453/2020\|EPI_ISL_578598\|202 | ···················· | ···················· | ····················· | ···················· | ···················· | ···················· |
| L | 8354 | hCoV-19/USA/WI-WSLH-200456/2020\|EPI_ISL_578599\|202 | ···················· | ···················· | ····················· | ···················· | ···················· | ···················· |
| L | 8355 | hCoV-19/England/QEUH-9E47C8/2020\|EPI_ISL_579856\|20 | ···················· | ···················· | ····················· | ···················· | ···T················ | ···················· |
| L | 8356 | hCoV-19/England/QEUH-9E5062/2020\|EPI_ISL_579857\|20 | ···················· | ·A·················· | ····················· | ···················· | ···················· | ···················· |
| L | 8357 | hCoV-19/England/QEUH-9E4BAE/2020\|EPI_ISL_579858\|20 | ···················· | ···················· | ····················· | ···················· | ···················· | ···················· |
| L | 8358 | hCoV-19/England/QEUH-9E4B80/2020\|EPI_ISL_579859\|20 | ···················· | ···················· | ····················· | ···················· | ···················· | ···················· |
| L | 8359 | hCoV-19/England/QEUH-9E5624/2020\|EPI_ISL_579860\|20 | ···················· | ···················· | ····················· | ···················· | ···················· | ···················· |
| L | 8360 | hCoV-19/England/QEUH-9E5475/2020\|EPI_ISL_579861\|20 | ···················· | ···················· | ····················· | ···················· | ···················· | ···················· |
| L | 8361 | hCoV-19/England/QEUH-9E4C41/2020\|EPI_ISL_579862\|20 | ···················· | ···················· | ····················· | ···················· | ···················· | ···················· |
| L | 8362 | hCoV-19/England/QEUH-9E48D4/2020\|EPI_ISL_579863\|20 | ···················· | ···················· | ····················· | ···················· | ···················· | ···················· |
| L | 8363 | hCoV-19/England/QEUH-9E4E69/2020\|EPI_ISL_579864\|20 | ···················· | ···················· | ····················· | ···················· | ···················· | ···················· |
| L | 8364 | hCoV-19/England/QEUH-9E530F/2020\|EPI_ISL_579865\|20 | ···················· | ·A·················· | ····················· | ···················· | ···················· | ···················· |
| L | 8365 | hCoV-19/England/QEUH-9E4616/2020\|EPI_ISL_579867\|20 | ···················· | ···················· | ····················· | ···················· | ···················· | ···················· |
| L | 8366 | hCoV-19/England/QEUH-9E49E0/2020\|EPI_ISL_579868\|20 | ···················· | ···················· | ····················· | ···················· | ···················· | ···················· |
| L | 8367 | hCoV-19/England/QEUH-9E4C14/2020\|EPI_ISL_579869\|20 | ···················· | ···················· | ····················· | ···················· | ···················· | ···················· |
| L | 8368 | hCoV-19/England/QEUH-9E4212/2020\|EPI_ISL_579870\|20 | ···················· | ···················· | ····················· | ···················· | ···················· | ···················· |
| L | 8369 | hCoV-19/England/QEUH-9E4E5A/2020\|EPI_ISL_579871\|20 | ···················· | ···················· | ····················· | ···················· | ···················· | ···················· |
| L | 8370 | hCoV-19/England/QEUH-9E4467/2020\|EPI_ISL_579872\|20 | ···················· | ···················· | ····················· | ···················· | ···················· | ···················· |
| L | 8371 | hCoV-19/England/QEUH-9E4BBD/2020\|EPI_ISL_579873\|20 | ···················· | ···················· | ····················· | ···················· | ···················· | ···················· |
| L | 8372 | hCoV-19/England/QEUH-9E4F48/2020\|EPI_ISL_579874\|20 | ···················· | ·A·················· | ····················· | ···················· | ···················· | ··A················· |
| L | 8373 | hCoV-19/England/QEUH-9E4F93/2020\|EPI_ISL_579875\|20 | ···················· | ···················· | ····················· | ···················· | ···················· | ···················· |
| L | 8374 | hCoV-19/England/QEUH-9E41BB/2020\|EPI_ISL_579876\|20 | ···················· | ···················· | ····················· | ···················· | ···················· | ···················· |
| L | 8375 | hCoV-19/England/QEUH-9E5484/2020\|EPI_ISL_579877\|20 | ···················· | ···················· | ····················· | ···················· | ···················· | ···················· |
| L | 8376 | hCoV-19/England/QEUH-9E5071/2020\|EPI_ISL_579878\|20 | ···················· | ···················· | ····················· | ···················· | ···················· | ···················· |
| L | 8377 | hCoV-19/England/QEUH-9E5545/2020\|EPI_ISL_579879\|20 | ···················· | ·A·················· | ····················· | ···················· | ···················· | ··A················· |
| L | 8378 | hCoV-19/England/QEUH-9E45FB/2020\|EPI_ISL_579880\|20 | ···················· | ···················· | ····················· | ···················· | ···················· | ···················· |
| L | 8379 | hCoV-19/England/QEUH-9E3E88/2020\|EPI_ISL_579881\|20 | ···················· | ···················· | ····················· | ···················· | ···················· | ···················· |
| L | 8380 | hCoV-19/England/QEUH-9E3F49/2020\|EPI_ISL_579882\|20 | ···················· | ···················· | ····················· | ···················· | ···················· | ···················· |
| L | 8381 | hCoV-19/England/QEUH-9E50DB/2020\|EPI_ISL_579883\|20 | ···················· | ···················· | ····················· | ···················· | ···················· | ···················· |
| L | 8382 | hCoV-19/England/QEUH-9E4FC0/2020\|EPI_ISL_579884\|20 | ···················· | ···················· | ····················· | ···················· | ···················· | ···················· |
| L | 8383 | hCoV-19/England/QEUH-9E4BCC/2020\|EPI_ISL_579885\|20 | ···················· | ···················· | ····················· | ···················· | ···················· | ···················· |
| L | 8384 | hCoV-19/England/QEUH-9E52B7/2020\|EPI_ISL_579886\|20 | ···················· | ···················· | ····················· | ···················· | ···················· | ···················· |
| L | 8385 | hCoV-19/England/QEUH-9E4B08/2020\|EPI_ISL_579887\|20 | ···················· | ···················· | ····················· | ···················· | ···················· | ···················· |
| L | 8386 | hCoV-19/England/QEUH-9E5035/2020\|EPI_ISL_579888\|20 | ···················· | ···················· | ····················· | ···················· | ···················· | ···················· |
| L | 8387 | hCoV-19/England/QEUH-9E4C9C/2020\|EPI_ISL_579889\|20 | ···················· | ···················· | ····················· | ···················· | ···················· | ···················· |
| L | 8388 | hCoV-19/England/QEUH-9E478C/2020\|EPI_ISL_579890\|20 | ···················· | ···················· | ····················· | ···················· | ···················· | ···················· |

L 8389 hCoV-19/England/QEUH-9E4B53/2020|EPI_ISL_579891|20 ···················· ···················· ···················· ···················· ···················· ····················
L 8390 hCoV-19/England/QEUH-9E41AC/2020|EPI_ISL_579892|20 ···················· ···················· ···················· ···················· ······T············· ····················
L 8391 hCoV-19/England/QEUH-9E532D/2020|EPI_ISL_579893|20 ···················· ···················· ···················· ···················· ···················· ····················
L 8392 hCoV-19/England/QEUH-9E3E79/2020|EPI_ISL_579894|20 ···················· ···················· ···················· ···················· ···················· ····················
L 8393 hCoV-19/England/QEUH-9E509F/2020|EPI_ISL_579895|20 ···················· ···················· ···················· ···················· ···················· ····················
L 8394 hCoV-19/England/QEUH-9E475F/2020|EPI_ISL_579896|20 ···················· ···················· ···················· ···················· ····T··············· ····················
L 8395 hCoV-19/England/QEUH-9E4FA2/2020|EPI_ISL_579897|20 ···················· ···················· ···················· ···················· ···················· ····················
L 8396 hCoV-19/England/QEUH-9E476E/2020|EPI_ISL_579898|20 ···················· ···················· ···················· ···················· ······T············· ····················
L 8397 hCoV-19/England/QEUH-9E477D/2020|EPI_ISL_579899|20 ···················· ···················· ···················· ···················· ···················· ····················
L 8398 hCoV-19/Wales/MILK-9EE940/2020|EPI_ISL_580841|2020 ···················· ···················· ···················· ···················· ···················· ····················
L 8399 hCoV-19/England/MILK-9F0468/2020|EPI_ISL_580842|20 ···················· ···················· ···················· ···················· ···················· ····················
L 8400 hCoV-19/England/MILK-9EEC1A/2020|EPI_ISL_580843|20 ···················· ···················· ···················· ···················· ···················· ····················
L 8401 hCoV-19/England/MILK-9EE7EC/2020|EPI_ISL_580844|20 ···················· ···················· ···················· ···················· ···················· ····················
L 8402 hCoV-19/England/MILK-9EEDF9/2020|EPI_ISL_580845|20 ···················· ···················· ···················· ···················· ···················· ····················
L 8403 hCoV-19/Wales/MILK-9F05DE/2020|EPI_ISL_580847|2020 ···················· ···················· ···················· ···················· ···················· ····················
L 8404 hCoV-19/Wales/MILK-9EEEC9/2020|EPI_ISL_580848|2020 ···················· ···················· ···················· ···················· ···················· ····················
L 8405 hCoV-19/England/ALDP-9EDAF3/2020|EPI_ISL_580849|20 ···················· ·A·················· ···················· ···················· ···················· ····················
L 8406 hCoV-19/Wales/MILK-9EEB3B/2020|EPI_ISL_580850|2020 ···················· ···················· ···················· ···················· ···················· ····················
L 8407 hCoV-19/England/ALDP-9EDA3F/2020|EPI_ISL_580851|20 ···················· ···················· ···················· ···················· ···················· ····················
L 8408 hCoV-19/England/MILK-9EE5B5/2020|EPI_ISL_580852|20 ···················· ···················· ···················· ···················· ···················· ····················
L 8409 hCoV-19/England/MILK-9EE6D0/2020|EPI_ISL_580854|20 ···················· ···················· ···················· ···················· ···················· ····················
L 8410 hCoV-19/Wales/MILK-9EECFC/2020|EPI_ISL_580856|2020 ···················· ···················· ···················· ···················· ···················· ····················
L 8411 hCoV-19/England/MILK-9EEE32/2020|EPI_ISL_580857|20 ···················· ···················· ···················· ···················· ···················· ····················
L 8412 hCoV-19/England/QEUH-9DC383/2020|EPI_ISL_580858|20 ···················· ···················· ···················· ···················· ···················· ····················
L 8413 hCoV-19/England/QEUH-9DC550/2020|EPI_ISL_580859|20 ···················· ···················· ···················· ···················· ···················· ····················
L 8414 hCoV-19/England/QEUH-9DCDD0/2020|EPI_ISL_580860|20 ···················· ···················· ···················· ···················· ···················· ····················
L 8415 hCoV-19/Wales/MILK-9E11DC/2020|EPI_ISL_580861|2020 ···················· ···················· ···················· ···················· ···················· ····················
L 8416 hCoV-19/England/QEUH-9DC67B/2020|EPI_ISL_580862|20 ···················· ···················· ···················· ···················· ···················· ····················
L 8417 hCoV-19/England/QEUH-9DCBB8/2020|EPI_ISL_580863|20 ···················· ·A·················· ···················· ···················· ···················· ···A················
L 8418 hCoV-19/Wales/MILK-9E0F3D/2020|EPI_ISL_580864|2020 ···················· ···················· ···················· ···················· ···················· ····················
L 8419 hCoV-19/England/QEUH-9DB0E7/2020|EPI_ISL_580865|20 ···················· ···················· ···················· ···················· ···················· ····················
L 8420 hCoV-19/England/QEUH-9DAFFA/2020|EPI_ISL_580866|20 ···················· ···················· ···················· ···················· ···················· ····················
L 8421 hCoV-19/England/QEUH-9DACE4/2020|EPI_ISL_580867|20 ···················· ···················· ···················· ···················· ···················· ····················
L 8422 hCoV-19/England/QEUH-9DC972/2020|EPI_ISL_580868|20 ···················· ···················· ···················· ···················· ···················· ····················
L 8423 hCoV-19/England/QEUH-9DAACC/2020|EPI_ISL_580869|20 ···················· ···················· ···················· ···················· ···················· ····················
L 8424 hCoV-19/England/QEUH-9DC954/2020|EPI_ISL_580871|20 ···················· ·A·················· ···················· ···················· ···················· ···A················
L 8425 hCoV-19/England/QEUH-9DCC6A/2020|EPI_ISL_580872|20 ···················· ···················· ···················· ···················· ···················· ····················
L 8426 hCoV-19/Wales/MILK-9E0FE2/2020|EPI_ISL_580873|2020 ···················· ···················· ···················· ···················· ···················· ····················
L 8427 hCoV-19/England/QEUH-9DCAAC/2020|EPI_ISL_580874|20 ···················· ···················· ···················· ···················· ···················· ····················
L 8428 hCoV-19/Wales/MILK-9E11EB/2020|EPI_ISL_580875|2020 ···················· ···················· ···················· ···················· ···················· ····················
L 8429 hCoV-19/Wales/MILK-9E1181/2020|EPI_ISL_580877|2020 ···················· ···················· ···················· ···················· ···················· ····················
L 8430 hCoV-19/England/QEUH-9DC620/2020|EPI_ISL_580878|20 ···················· ···················· ···················· ···················· ···················· ····················
L 8431 hCoV-19/England/QEUH-9DB0AB/2020|EPI_ISL_580879|20 ···················· ···················· ···················· ···················· ···················· ····················
L 8432 hCoV-19/England/QEUH-9DC9AF/2020|EPI_ISL_580880|20 ···················· ·A·················· ···················· ···················· ···················· ···A················
L 8433 hCoV-19/England/QEUH-9DAEFD/2020|EPI_ISL_580881|20 ···················· ···················· ···················· ···················· ····T··············· ····················
L 8434 hCoV-19/Wales/MILK-9E0E12/2020|EPI_ISL_580882|2020 ···················· ···················· ···················· ···················· ···················· ····················
L 8435 hCoV-19/England/QEUH-9DAC20/2020|EPI_ISL_580883|20 ···················· ···················· ···················· ···················· ···················· ····················
L 8436 hCoV-19/England/QEUH-9DAFCD/2020|EPI_ISL_580884|20 ···················· ···················· ···················· ···················· ···················· ····················
L 8437 hCoV-19/England/MILK-9E0D9D/2020|EPI_ISL_580885|20 ···················· ···················· ···················· ···················· ······T············· ····················
L 8438 hCoV-19/England/QEUH-9DD346/2020|EPI_ISL_580886|20 ···················· ···················· ···················· ···················· ···················· ····················
L 8439 hCoV-19/England/QEUH-9DD2C1/2020|EPI_ISL_580887|20 ···················· ···················· ···················· ···················· ···················· ····················
L 8440 hCoV-19/England/QEUH-9DD355/2020|EPI_ISL_580888|20 ···················· ···················· ···················· ···················· ···················· ····················
L 8441 hCoV-19/England/QEUH-9DAEDF/2020|EPI_ISL_580889|20 ···················· ···················· ···················· ···················· ···················· ····················
L 8442 hCoV-19/Wales/MILK-9E0DCA/2020|EPI_ISL_580896|2020 ···················· ···················· ···················· ···················· ···················· ····················
L 8443 hCoV-19/England/MILK-9E0DE8/2020|EPI_ISL_580897|20 ···················· ···················· ···················· ···················· ···················· ····················
L 8444 hCoV-19/USA/MI-UM-10035665108/2020|EPI_ISL_583250| ···················· ···················· ···················· ···················· ···················· ····················
L 8445 hCoV-19/USA/MI-UM-10035663652/2020|EPI_ISL_583251| ···················· ···················· ···················· ···················· ···················· ····················
L 8448 hCoV-19/USA/MI-UM-10035712935/2020|EPI_ISL_583254| ···················· ·A·················· ···················· ···················· ···················· ····················
L 8449 hCoV-19/USA/CA-QDX-514/2020|EPI_ISL_571177|2020-03 ···················· ···················· ···················· ···················· ···················· ····················
L 8450 hCoV-19/USA/CT-QDX-426/2020|EPI_ISL_571178|2020-03 ···················· ···················· ···················· ···················· ···················· ····················
L 8451 hCoV-19/USA/CT-QDX-427/2020|EPI_ISL_571179|2020-03 ···················· ···················· ···················· ···················· ···················· ····················
L 8452 hCoV-19/USA/CT-QDX-428/2020|EPI_ISL_571180|2020-03 ···················· ···················· ···················· ···················· ···················· ····················
L 8453 hCoV-19/USA/CT-QDX-429/2020|EPI_ISL_571181|2020-03 ···················· ···················· ···················· ···················· ···················· ····················
L 8454 hCoV-19/USA/FL-QDX-422/2020|EPI_ISL_571182|2020-03 ···················· ···················· ···················· ···················· ···················· ····················
L 8455 hCoV-19/USA/HI-QDX-411/2020|EPI_ISL_571183|2020-03 ···················· ···················· ···················· ···················· ···················· ····················
L 8456 hCoV-19/USA/HI-QDX-412/2020|EPI_ISL_571184|2020-03 ···················· ···················· ···················· ···················· ···················· ····················
L 8457 hCoV-19/USA/HI-QDX-424/2020|EPI_ISL_571185|2020-03 ···················· ···················· ···················· ···················· ···················· ····················
L 8458 hCoV-19/USA/IL-QDX-431/2020|EPI_ISL_571186|2020-03 ···················· ···················· ···················· ···················· ···················· ····················
L 8459 hCoV-19/USA/LA-QDX-410/2020|EPI_ISL_571187|2020-03 ···················· ···················· ···················· ···················· ···················· ····················
L 8460 hCoV-19/USA/LA-QDX-416/2020|EPI_ISL_571188|2020-03 ···················· ···················· ···················· ···················· ···················· ····················
L 8461 hCoV-19/USA/MI-QDX-454/2020|EPI_ISL_571189|2020-03 ···················· ···················· ···················· ···················· ···················· ····················
L 8462 hCoV-19/USA/MI-QDX-455/2020|EPI_ISL_571190|2020-03 ···················· ···················· ···················· ···················· ···················· ····················
L 8463 hCoV-19/USA/MI-QDX-456/2020|EPI_ISL_571191|2020-03 ···················· ···················· ···················· ···················· ···················· ····················
L 8464 hCoV-19/USA/MI-QDX-457/2020|EPI_ISL_571192|2020-03 ···················· ···················· ···················· ···················· ···················· ····················
L 8465 hCoV-19/USA/MI-QDX-458/2020|EPI_ISL_571193|2020-03 ···················· ···················· ···················· ···················· ···················· ····················
L 8466 hCoV-19/USA/MI-QDX-459/2020|EPI_ISL_571194|2020-03 ···················· ···················· ···················· ···················· ···················· ····················
L 8467 hCoV-19/USA/MI-QDX-460/2020|EPI_ISL_571195|2020-03 ···················· ···················· ···················· ···················· ···················· ····················
L 8468 hCoV-19/USA/MI-QDX-461/2020|EPI_ISL_571196|2020-03 ···················· ···················· ···················· ···················· ···················· ····················
L 8469 hCoV-19/USA/MI-QDX-462/2020|EPI_ISL_571197|2020-03 ···················· ···················· ···················· ···················· ···················· ····················
L 8470 hCoV-19/USA/MI-QDX-463/2020|EPI_ISL_571198|2020-03 ···················· ···················· ···················· ···················· ···················· ····················

| | | | | | | | | |
|---|---|---|---|---|---|---|---|---|
| L | 8471 | hCoV-19/USA/MI-QDX-464/2020\|EPI_ISL_571199\|2020-03 | ···················· | ···················· | ····················· | ···················· | ···················· | ···················· |
| L | 8472 | hCoV-19/Scotland/CVR4314/2020\|EPI_ISL_573771\|2020- | ···················· | ···················· | ····················· | ···················· | ··············N····· | N··················· |
| L | 8473 | hCoV-19/England/QEUH-9AE9B5/2020\|EPI_ISL_577260\|20 | ···················· | ···················· | ····················· | ···················· | ···················· | ···················· |
| L | 8474 | hCoV-19/England/QEUH-9AE9C4/2020\|EPI_ISL_577261\|20 | ···················· | ···················· | ····················· | ···················· | ···················· | ···················· |
| L | 8475 | hCoV-19/England/QEUH-9AE9D3/2020\|EPI_ISL_577262\|20 | ···················· | ···················· | ····················· | ···················· | ···················· | ···················· |
| L | 8476 | hCoV-19/England/QEUH-9AE9E2/2020\|EPI_ISL_577263\|20 | ···················· | ···················· | ····················· | ···················· | ···················· | ···················· |
| L | 8477 | hCoV-19/England/QEUH-9AEA0D/2020\|EPI_ISL_577264\|20 | ···················· | ·A·················· | ····················· | ···················· | ···················· | ···················· |
| L | 8478 | hCoV-19/England/QEUH-9AEA2B/2020\|EPI_ISL_577266\|20 | ···················· | ···················· | ····················· | ···················· | ···················· | ···················· |
| L | 8479 | hCoV-19/England/QEUH-9D3128/2020\|EPI_ISL_577310\|20 | ···················· | ···················· | ····················· | ···················· | ···················· | ···················· |
| L | 8481 | hCoV-19/USA/WI-WSLH-200444/2020\|EPI_ISL_578591\|202 | ···················· | ···················· | ····················· | ···················· | ···················· | ···················· |
| L | 8483 | hCoV-19/USA/WI-WSLH-200448/2020\|EPI_ISL_578593\|202 | ···················· | ···················· | ····················· | ···················· | ···················· | ···················· |
| L | 8485 | hCoV-19/USA/WI-WSLH-200450/2020\|EPI_ISL_578595\|202 | ···················· | ···················· | ····················· | ···················· | ···················· | ···················· |
| L | 8487 | hCoV-19/USA/WI-WSLH-200452/2020\|EPI_ISL_578597\|202 | ···················· | ···················· | ····················· | ···················· | ···················· | ···················· |
| L | 8488 | hCoV-19/USA/WI-WSLH-200458/2020\|EPI_ISL_578600\|202 | ···················· | ···················· | ····················· | ···················· | ···················· | ···················· |
| L | 8489 | hCoV-19/USA/WI-WSLH-200459/2020\|EPI_ISL_578601\|202 | ···················· | ···················· | ····················· | ···················· | ···················· | ···················· |
| L | 8490 | hCoV-19/USA/WI-WSLH-200460/2020\|EPI_ISL_578602\|202 | ···················· | ···················· | ····················· | ···················· | ···················· | ···················· |
| L | 8491 | hCoV-19/USA/WI-WSLH-200461/2020\|EPI_ISL_578603\|202 | ···················· | ···················· | ····················· | ···················· | ···················· | ···················· |
| L | 8492 | hCoV-19/USA/WI-WSLH-200462/2020\|EPI_ISL_578604\|202 | ···················· | ···················· | ····················· | ···················· | ···················· | ···················· |
| L | 8493 | hCoV-19/USA/WI-WSLH-200463/2020\|EPI_ISL_578605\|202 | ···················· | ···················· | ····················· | ···················· | ···················· | ···················· |
| L | 8494 | hCoV-19/USA/WI-WSLH-200464/2020\|EPI_ISL_578606\|202 | ···················· | ···················· | ····················· | ···················· | ···················· | ···················· |
| L | 8495 | hCoV-19/USA/WI-WSLH-200466/2020\|EPI_ISL_578608\|202 | ···················· | ···················· | ····················· | ···················· | ···················· | ···················· |
| L | 8496 | hCoV-19/USA/WI-WSLH-200467/2020\|EPI_ISL_578609\|202 | ···················· | ···················· | ····················· | ···················· | ···················· | ···················· |
| L | 8497 | hCoV-19/USA/WI-WSLH-200468/2020\|EPI_ISL_578610\|202 | ···················· | ···················· | ····················· | ···················· | ···················· | ···················· |
| L | 8498 | hCoV-19/USA/WI-WSLH-200470/2020\|EPI_ISL_578611\|202 | ···················· | ···················· | ····················· | ···················· | ···················· | ···················· |
| L | 8500 | hCoV-19/USA/WI-WSLH-200472/2020\|EPI_ISL_578613\|202 | ···················· | ···················· | ····················· | ···················· | ···················· | ···················· |
| L | 8501 | hCoV-19/USA/WI-WSLH-200473/2020\|EPI_ISL_578614\|202 | ···················· | ···················· | ····················· | ···················· | ···················· | ···················· |
| L | 8502 | hCoV-19/USA/WI-WSLH-200474/2020\|EPI_ISL_578615\|202 | ···················· | ···················· | ····················· | ···················· | ···················· | ···················· |
| L | 8503 | hCoV-19/USA/WI-WSLH-200475/2020\|EPI_ISL_578616\|202 | ···················· | ···················· | ····················· | ···················· | ···················· | ···················· |
| L | 8506 | hCoV-19/USA/WI-WSLH-200478/2020\|EPI_ISL_578619\|202 | ···················· | ···················· | ····················· | ···················· | ···················· | ···················· |
| L | 8513 | hCoV-19/USA/WI-WSLH-200485/2020\|EPI_ISL_578626\|202 | ···················· | ···················· | ····················· | ···················· | ···················· | ···················· |
| L | 8514 | hCoV-19/USA/WI-WSLH-200486/2020\|EPI_ISL_578627\|202 | ···················· | ···················· | ····················· | ···················· | ···················· | ···················· |
| L | 8517 | hCoV-19/USA/WI-WSLH-200489/2020\|EPI_ISL_578630\|202 | ···················· | ···················· | ····················· | ···················· | ···················· | ···················· |
| L | 8521 | hCoV-19/USA/WI-WSLH-200493/2020\|EPI_ISL_578634\|202 | ···················· | ···················· | ····················· | ···················· | ···················· | ···················· |
| L | 8527 | hCoV-19/USA/WI-WSLH-200499/2020\|EPI_ISL_578640\|202 | ···················· | ···················· | ····················· | ···················· | ···················· | ···················· |
| L | 8528 | hCoV-19/USA/WI-WSLH-200500/2020\|EPI_ISL_578641\|202 | ···················· | ···················· | ····················· | ···················· | ···················· | ···················· |
| L | 8533 | hCoV-19/England/QEUH-9E4AFC/2020\|EPI_ISL_579900\|20 | ···················· | ···················· | ····················· | ···················· | ···················· | ···················· |
| L | 8534 | hCoV-19/England/QEUH-9E4B44/2020\|EPI_ISL_579901\|20 | ···················· | ···················· | ····················· | ···················· | ···················· | ···················· |
| L | 8535 | hCoV-19/England/QEUH-9E528A/2020\|EPI_ISL_579902\|20 | ···················· | ···················· | ····················· | ···················· | ···················· | ···················· |
| L | 8536 | hCoV-19/England/QEUH-9E4DE4/2020\|EPI_ISL_579903\|20 | ···················· | ·A·················· | ····················· | ···················· | ···················· | ···················· |
| L | 8537 | hCoV-19/England/QEUH-9E49C2/2020\|EPI_ISL_579904\|20 | ···················· | ···················· | ····················· | ···················· | ···················· | ···················· |
| L | 8538 | hCoV-19/England/QEUH-9E5536/2020\|EPI_ISL_579905\|20 | ···················· | ···················· | ····················· | ···················· | ···················· | ···················· |
| L | 8539 | hCoV-19/England/QEUH-9E5080/2020\|EPI_ISL_579906\|20 | ···················· | ···················· | ····················· | ···················· | ···················· | ···················· |
| L | 8540 | hCoV-19/England/QEUH-9E4995/2020\|EPI_ISL_579908\|20 | ···················· | ···················· | ····················· | ···················· | ···················· | ···················· |
| L | 8541 | hCoV-19/England/QEUH-9E4221/2020\|EPI_ISL_579909\|20 | ···················· | ·A·················· | ····················· | ···················· | ···················· | ···A················ |
| L | 8542 | hCoV-19/England/QEUH-9E5563/2020\|EPI_ISL_579910\|20 | ···················· | ···················· | ····················· | ···················· | ···················· | ···················· |
| L | 8543 | hCoV-19/England/QEUH-9E43C4/2020\|EPI_ISL_579911\|20 | ···················· | ···················· | ····················· | ···················· | ···················· | ···················· |
| L | 8544 | hCoV-19/England/QEUH-9E493B/2020\|EPI_ISL_579912\|20 | ···················· | ···················· | ····················· | ···················· | ···················· | ···················· |
| L | 8545 | hCoV-19/England/QEUH-9E4DB7/2020\|EPI_ISL_579913\|20 | ···················· | ···················· | ····················· | ···················· | ···················· | ···················· |
| L | 8546 | hCoV-19/England/QEUH-9E4379/2020\|EPI_ISL_579914\|20 | ···················· | ····A··············· | ····················· | ···················· | ···················· | ···················· |
| L | 8547 | hCoV-19/England/QEUH-9E4476/2020\|EPI_ISL_579915\|20 | ···················· | ···················· | ····················· | ···················· | ···················· | ···················· |
| L | 8548 | hCoV-19/England/QEUH-9E418E/2020\|EPI_ISL_579916\|20 | ···················· | ···················· | ····················· | ···················· | ···················· | ···················· |
| L | 8549 | hCoV-19/England/QEUH-9E50BD/2020\|EPI_ISL_579917\|20 | ···················· | ···················· | ····················· | ···················· | ···················· | ···················· |
| L | 8550 | hCoV-19/England/ALDP-9EA012/2020\|EPI_ISL_579919\|20 | ···················· | ···················· | ····················· | ···················· | ···················· | ···················· |
| L | 8551 | hCoV-19/England/ALDP-9E9BA9/2020\|EPI_ISL_579921\|20 | ···················· | ···················· | ····················· | ···················· | ···················· | ···················· |
| L | 8552 | hCoV-19/England/ALDP-9EA883/2020\|EPI_ISL_579922\|20 | ···················· | ···················· | ····················· | ···················· | ···················· | ···················· |
| L | 8553 | hCoV-19/England/ALDP-9EA06D/2020\|EPI_ISL_579923\|20 | ···················· | ···················· | ····················· | ···················· | ···················· | ···················· |
| L | 8554 | hCoV-19/England/ALDP-9EA5E6/2020\|EPI_ISL_579924\|20 | ···················· | ···················· | ····················· | ···················· | ···················· | ···················· |
| L | 8555 | hCoV-19/England/ALDP-9EAF8E/2020\|EPI_ISL_579925\|20 | ···················· | ···················· | ····················· | ···················· | ···················· | ···················· |
| L | 8556 | hCoV-19/England/ALDP-9E9D85/2020\|EPI_ISL_579927\|20 | ···················· | ···················· | ····················· | ···················· | ···················· | ···················· |
| L | 8557 | hCoV-19/England/ALDP-9EA777/2020\|EPI_ISL_579928\|20 | ···················· | ···················· | ····················· | ···················· | ···················· | ···················· |
| L | 8558 | hCoV-19/England/ALDP-9EA829/2020\|EPI_ISL_579929\|20 | ···················· | ···················· | ····················· | ···················· | ···················· | ···················· |
| L | 8559 | hCoV-19/England/ALDP-9EA531/2020\|EPI_ISL_579930\|20 | ···················· | ···················· | ····················· | ···················· | ···················· | ···················· |
| L | 8560 | hCoV-19/England/ALDP-9EA6E3/2020\|EPI_ISL_579931\|20 | ···················· | ···················· | ····················· | ···················· | ···················· | ···················· |
| L | 8561 | hCoV-19/England/ALDP-9EAD39/2020\|EPI_ISL_579932\|20 | ···················· | ···················· | ····················· | ···················· | ···················· | ···················· |
| L | 8562 | hCoV-19/England/ALDP-9EAD93/2020\|EPI_ISL_579933\|20 | ···················· | ···················· | ····················· | ···················· | ···················· | ···················· |
| L | 8563 | hCoV-19/England/ALDP-9EA2FE/2020\|EPI_ISL_579934\|20 | ···················· | ···················· | ····················· | ···················· | ···················· | ···················· |
| L | 8564 | hCoV-19/England/ALDP-9EA15B/2020\|EPI_ISL_579935\|20 | ···················· | ···················· | ····················· | ···················· | ···················· | ···················· |
| L | 8565 | hCoV-19/England/ALDP-9E79DE/2020\|EPI_ISL_579936\|20 | ···················· | ···················· | ····················· | ···················· | ···················· | ···················· |
| L | 8566 | hCoV-19/England/ALDP-9E9B12/2020\|EPI_ISL_579937\|20 | ···················· | ···················· | ····················· | ···················· | ···················· | ···················· |
| L | 8567 | hCoV-19/England/ALDP-9E9B9A/2020\|EPI_ISL_579938\|20 | ···················· | ···················· | ····················· | ···················· | ···················· | ···················· |
| L | 8568 | hCoV-19/England/ALDP-9EA5F5/2020\|EPI_ISL_579940\|20 | ···················· | ···················· | ····················· | ···················· | ···················· | ···················· |
| L | 8569 | hCoV-19/England/ALDP-9EA416/2020\|EPI_ISL_579941\|20 | ···················· | ···················· | ····················· | ···················· | ···················· | ···················· |
| L | 8570 | hCoV-19/England/ALDP-9EA355/2020\|EPI_ISL_579942\|20 | ···················· | ···················· | ····················· | ···················· | ···················· | ···················· |
| L | 8571 | hCoV-19/England/ALDP-9E79B0/2020\|EPI_ISL_579943\|20 | ···················· | ···················· | ····················· | ···················· | ···················· | ···················· |
| L | 8572 | hCoV-19/England/ALDP-9EAFE8/2020\|EPI_ISL_579944\|20 | ···················· | ···················· | ····················· | ···················· | ···················· | ···················· |
| L | 8573 | hCoV-19/England/ALDP-9EAFBB/2020\|EPI_ISL_579945\|20 | ···················· | ···················· | ····················· | ···················· | ···················· | ···················· |
| L | 8574 | hCoV-19/England/ALDP-9EAB4E/2020\|EPI_ISL_579946\|20 | ···················· | ···················· | ····················· | ···················· | ···················· | ···················· |
| L | 8575 | hCoV-19/England/ALDP-9EA522/2020\|EPI_ISL_579947\|20 | ···················· | ···················· | ····················· | ···················· | ···················· | ···················· |
| L | 8576 | hCoV-19/England/ALDP-9EAB20/2020\|EPI_ISL_579948\|20 | ···················· | ···················· | ····················· | ···················· | ···················· | ···················· |
| L | 8577 | hCoV-19/England/ALDP-9EAF42/2020\|EPI_ISL_579950\|20 | ···················· | ···················· | ····················· | ···················· | ···················· | ···················· |

| | | | | | | | | |
|---|---|---|---|---|---|---|---|---|
| L | 8578 | hCoV-19/England/ALDP-9EA689/2020\|EPI_ISL_579952\|20 | ···················· | ···················· | ····················· | ···················· | ···················· | ···················· |
| L | 8579 | hCoV-19/England/ALDP-9EA892/2020\|EPI_ISL_579954\|20 | ···················· | ···················· | ····················· | ···················· | ···················· | ···················· |
| L | 8580 | hCoV-19/England/ALDP-9E9F43/2020\|EPI_ISL_579955\|20 | ···················· | ···················· | ····················· | ···················· | ···················· | ···················· |
| L | 8581 | hCoV-19/England/ALDP-9EA346/2020\|EPI_ISL_579956\|20 | ···················· | ···················· | ····················· | ···················· | ···················· | ···················· |
| L | 8582 | hCoV-19/England/ALDP-9EAE18/2020\|EPI_ISL_579957\|20 | ···················· | ···················· | ····················· | ···················· | ···················· | ···················· |
| L | 8583 | hCoV-19/England/ALDP-9EA030/2020\|EPI_ISL_579958\|20 | ···················· | ···················· | ····················· | ···················· | ···················· | ···················· |
| L | 8584 | hCoV-19/England/ALDP-9EACE1/2020\|EPI_ISL_579959\|20 | ···················· | ···················· | ····················· | ···················· | ···················· | ···················· |
| L | 8585 | hCoV-19/England/ALDP-9EA3A0/2020\|EPI_ISL_579961\|20 | ···················· | ···················· | ····················· | ···················· | ···················· | ···················· |
| L | 8586 | hCoV-19/England/ALDP-9EAC4B/2020\|EPI_ISL_579962\|20 | ···················· | ···················· | ····················· | ···················· | ···················· | ···················· |
| L | 8587 | hCoV-19/England/ALDP-9EAA14/2020\|EPI_ISL_579963\|20 | ···················· | ···················· | ····················· | ···················· | ···················· | ···················· |
| L | 8588 | hCoV-19/England/ALDP-9EAEEB/2020\|EPI_ISL_579964\|20 | ···················· | ···················· | ····················· | ···················· | ···················· | ···················· |
| L | 8589 | hCoV-19/England/ALDP-9EA197/2020\|EPI_ISL_579965\|20 | ···················· | ···················· | ····················· | ···················· | ···················· | ···················· |
| L | 8590 | hCoV-19/England/ALDP-9EA443/2020\|EPI_ISL_579966\|20 | ···················· | ···················· | ····················· | ···················· | ···················· | ···················· |
| L | 8591 | hCoV-19/England/ALDP-9EB05D/2020\|EPI_ISL_579967\|20 | ···················· | ···················· | ····················· | ···················· | ···················· | ···················· |
| L | 8592 | hCoV-19/England/ALDP-9EA3CE/2020\|EPI_ISL_579968\|20 | ···················· | ···················· | ····················· | ···················· | ···················· | ···················· |
| L | 8593 | hCoV-19/England/ALDP-9EA874/2020\|EPI_ISL_579969\|20 | ···················· | ···················· | ····················· | ···················· | ···················· | ···················· |
| L | 8594 | hCoV-19/England/ALDP-9E9C2E/2020\|EPI_ISL_579970\|20 | ···················· | ···················· | ····················· | ···················· | ···················· | ···················· |
| L | 8595 | hCoV-19/England/ALDP-9E9AF7/2020\|EPI_ISL_579972\|20 | ···················· | ····A··············· | ····················· | ···················· | ···················· | ···················· |
| L | 8596 | hCoV-19/England/ALDP-9EAE09/2020\|EPI_ISL_579973\|20 | ···················· | ···················· | ····················· | ···················· | ···················· | ···················· |
| L | 8597 | hCoV-19/England/ALDP-9EB04E/2020\|EPI_ISL_579974\|20 | ···················· | ···················· | ····················· | ···················· | ···················· | ···················· |
| L | 8598 | hCoV-19/England/ALDP-9E9FF8/2020\|EPI_ISL_579975\|20 | ···················· | ···················· | ····················· | ···················· | ···················· | ···················· |
| L | 8599 | hCoV-19/England/ALDP-9E9C88/2020\|EPI_ISL_579976\|20 | ···················· | ···················· | ····················· | ···················· | ···················· | ···················· |
| L | 8600 | hCoV-19/England/QEUH-9DCA8E/2020\|EPI_ISL_580891\|20 | ···················· | ···················· | ····················· | ···················· | ···················· | ···················· |
| L | 8601 | hCoV-19/England/MILK-9E1118/2020\|EPI_ISL_580892\|20 | ···················· | ···················· | ····················· | ···················· | ···················· | ···················· |
| L | 8602 | hCoV-19/England/QEUH-9DC198/2020\|EPI_ISL_580893\|20 | ···················· | ···················· | ····················· | ···················· | ···················· | ···················· |
| L | 8603 | hCoV-19/England/QEUH-9DCEEC/2020\|EPI_ISL_580894\|20 | ···················· | ···················· | ····················· | ···················· | ···················· | ···················· |
| L | 8604 | hCoV-19/Wales/MILK-9E0F4C/2020\|EPI_ISL_580898\|2020 | ···················· | ···················· | ····················· | ···················· | ···················· | ···················· |
| L | 8605 | hCoV-19/England/QEUH-9DC68A/2020\|EPI_ISL_580899\|20 | ···················· | ···················· | ····················· | ···················· | ···················· | ···················· |
| L | 8606 | hCoV-19/Wales/MILK-9E0D8E/2020\|EPI_ISL_580900\|2020 | ···················· | ···················· | ····················· | ···················· | ···················· | ···················· |
| L | 8607 | hCoV-19/England/QEUH-9DC5C9/2020\|EPI_ISL_580901\|20 | ···················· | ·A·················· | ····················· | ···················· | ···················· | ···A················ |
| L | 8608 | hCoV-19/Wales/MILK-9E12BB/2020\|EPI_ISL_580902\|2020 | ···················· | ···················· | ····················· | ···················· | ···················· | ···················· |
| L | 8609 | hCoV-19/England/MILK-9E10EE/2020\|EPI_ISL_580903\|20 | ···················· | ···················· | ····················· | ···················· | ···················· | ···················· |
| L | 8610 | hCoV-19/Wales/MILK-9E1260/2020\|EPI_ISL_580904\|2020 | ···················· | ···················· | ····················· | ···················· | ···················· | ···················· |
| L | 8611 | hCoV-19/England/QEUH-9DC611/2020\|EPI_ISL_580905\|20 | ···················· | ·A·················· | ····················· | ···················· | ···················· | ···A················ |
| L | 8612 | hCoV-19/England/QEUH-9DCB5E/2020\|EPI_ISL_580906\|20 | ···················· | ···················· | ····················· | ···················· | ···················· | ···················· |
| L | 8613 | hCoV-19/England/QEUH-9DAE0C/2020\|EPI_ISL_580907\|20 | ···················· | ···················· | ····················· | ···················· | ···················· | ···················· |
| L | 8614 | hCoV-19/England/QEUH-9DAFAF/2020\|EPI_ISL_580908\|20 | ···················· | ···················· | ····················· | ···················· | ···················· | ···················· |
| L | 8615 | hCoV-19/England/MILK-9E1084/2020\|EPI_ISL_580910\|20 | ···················· | ···················· | ····················· | ···················· | ···················· | ···················· |
| L | 8616 | hCoV-19/England/QEUH-9DC6A8/2020\|EPI_ISL_580911\|20 | ···················· | ···················· | ····················· | ···················· | ···················· | ···················· |
| L | 8617 | hCoV-19/England/QEUH-9DCEDD/2020\|EPI_ISL_580912\|20 | ···················· | ···················· | ····················· | ···················· | ···················· | ···················· |
| L | 8618 | hCoV-19/England/MILK-9E1066/2020\|EPI_ISL_580913\|20 | ···················· | ···················· | ····················· | ···················· | ···················· | ···················· |
| L | 8619 | hCoV-19/England/QEUH-9DC963/2020\|EPI_ISL_580914\|20 | ···················· | ···················· | ····················· | ···················· | ···················· | ···················· |
| L | 8620 | hCoV-19/England/MILK-9E0EB8/2020\|EPI_ISL_580915\|20 | ···················· | ···················· | ····················· | ···················· | ···················· | ···················· |
| L | 8621 | hCoV-19/England/QEUH-9DC365/2020\|EPI_ISL_580916\|20 | ···················· | ·A·················· | ····················· | ···················· | ···················· | ···A················ |
| L | 8622 | hCoV-19/England/MILK-9E10C0/2020\|EPI_ISL_580917\|20 | ···················· | ·A·················· | ····················· | ···················· | ···················· | ···················· |
| L | 8623 | hCoV-19/England/MILK-9E1136/2020\|EPI_ISL_580918\|20 | ···················· | ···················· | ····················· | ···················· | ···················· | ···················· |
| L | 8624 | hCoV-19/England/QEUH-9DC8C0/2020\|EPI_ISL_580919\|20 | ···················· | ···················· | ····················· | ···················· | ···················· | ···················· |
| L | 8625 | hCoV-19/England/QEUH-9DAB8D/2020\|EPI_ISL_580920\|20 | ···················· | ···················· | ····················· | ···················· | ···················· | ···················· |
| L | 8626 | hCoV-19/Wales/MILK-9E0F88/2020\|EPI_ISL_580921\|2020 | ···················· | ···················· | ····················· | ···················· | ···················· | ···················· |
| L | 8627 | hCoV-19/England/QEUH-9DC990/2020\|EPI_ISL_580922\|20 | ···················· | ···················· | ····················· | ···················· | ···················· | ···················· |
| L | 8628 | hCoV-19/England/QEUH-9DD2D0/2020\|EPI_ISL_580923\|20 | ···················· | ···················· | ····················· | ···················· | ···················· | ···················· |
| L | 8629 | hCoV-19/England/QEUH-9DCB4F/2020\|EPI_ISL_580924\|20 | ···················· | ·A·················· | ····················· | ···················· | ···················· | ···A················ |
| L | 8630 | hCoV-19/England/QEUH-9DC64E/2020\|EPI_ISL_580925\|20 | ···················· | ···················· | ····················· | ···················· | ···················· | ···················· |
| L | 8631 | hCoV-19/England/QEUH-9DCA60/2020\|EPI_ISL_580926\|20 | ···················· | ···················· | ····················· | ···················· | ······T············· | ···················· |
| L | 8632 | hCoV-19/England/QEUH-9DD328/2020\|EPI_ISL_580927\|20 | ···················· | ···················· | ····················· | ···················· | ···················· | ···················· |
| L | 8633 | hCoV-19/Wales/MILK-9E0FB5/2020\|EPI_ISL_580929\|2020 | ···················· | ···················· | ····················· | ···················· | ···················· | ···················· |
| L | 8634 | hCoV-19/England/QEUH-9DC866/2020\|EPI_ISL_580930\|20 | ···················· | ···················· | ····················· | ···················· | ···················· | ···················· |
| L | 8635 | hCoV-19/England/QEUH-9DC22C/2020\|EPI_ISL_580931\|20 | ···················· | ···················· | ····················· | ···················· | ···················· | ···················· |
| L | 8636 | hCoV-19/England/QEUH-9DC374/2020\|EPI_ISL_580932\|20 | ···················· | ·A·················· | ····················· | ···················· | ···················· | ···A················ |
| L | 8637 | hCoV-19/England/QEUH-9DC63F/2020\|EPI_ISL_580933\|20 | ···················· | ···················· | ····················· | ···················· | ···················· | ···················· |
| L | 8638 | hCoV-19/England/QEUH-9DC6D5/2020\|EPI_ISL_580934\|20 | ···················· | ···················· | ····················· | ···················· | ···················· | ···················· |
| L | 8639 | hCoV-19/Wales/MILK-9E1163/2020\|EPI_ISL_580935\|2020 | ···················· | ···················· | ····················· | ···················· | ···················· | ···················· |
| L | 8640 | hCoV-19/England/QEUH-9DAF72/2020\|EPI_ISL_580936\|20 | ···················· | ···················· | ····················· | ···················· | ···················· | ···················· |
| L | 8641 | hCoV-19/England/QEUH-9DAC11/2020\|EPI_ISL_580937\|20 | ···················· | ···················· | ····················· | ···················· | ···················· | ···················· |
| L | 8642 | hCoV-19/England/QEUH-9DC59C/2020\|EPI_ISL_580938\|20 | ···················· | ···················· | ····················· | ···················· | ···················· | ···················· |
| L | 8643 | hCoV-19/England/QEUH-9DC6B7/2020\|EPI_ISL_580939\|20 | ···················· | ···················· | ····················· | ···················· | ···················· | ···················· |
| L | 8644 | hCoV-19/England/MILK-9E0E9A/2020\|EPI_ISL_580940\|20 | ···················· | ···················· | ····················· | ···················· | ······T············· | ···················· |
| L | 8645 | hCoV-19/England/QEUH-9DCB7C/2020\|EPI_ISL_580941\|20 | ···················· | ···················· | ····················· | ···················· | ···················· | ···················· |
| L | 8646 | hCoV-19/England/QEUH-9DC541/2020\|EPI_ISL_580942\|20 | ···················· | ···················· | ····················· | ···················· | ···················· | ···················· |
| L | 8647 | hCoV-19/England/QEUH-9DADD2/2020\|EPI_ISL_580943\|20 | ···················· | ···················· | ····················· | ···················· | ···················· | ···················· |
| L | 8648 | hCoV-19/England/QEUH-9DADF0/2020\|EPI_ISL_580944\|20 | ···················· | ···················· | ····················· | ···················· | ···················· | ···················· |
| L | 8649 | hCoV-19/England/QEUH-9DCC88/2020\|EPI_ISL_580945\|20 | ···················· | ·A·················· | ····················· | ···················· | ···················· | ···················· |
| L | 8650 | hCoV-19/England/QEUH-9DCE19/2020\|EPI_ISL_580946\|20 | ···················· | ···················· | ····················· | ···················· | ···················· | ···················· |
| L | 8651 | hCoV-19/England/QEUH-9DCA7F/2020\|EPI_ISL_580947\|20 | ···················· | ···················· | ····················· | ···················· | ···················· | ···················· |
| L | 8652 | hCoV-19/Wales/MILK-9E0F5B/2020\|EPI_ISL_580948\|2020 | ···················· | ···················· | ····················· | ···················· | ···················· | ···················· |
| L | 8653 | hCoV-19/England/QEUH-9DABBA/2020\|EPI_ISL_580949\|20 | ···················· | ···················· | ····················· | ···················· | ···················· | ···················· |
| L | 8654 | hCoV-19/England/MILK-9E1093/2020\|EPI_ISL_580950\|20 | ···················· | ···················· | ····················· | ···················· | ···················· | ···················· |
| L | 8655 | hCoV-19/England/MILK-9E13A9/2020\|EPI_ISL_580951\|20 | ···················· | ···················· | ····················· | ···················· | ···················· | ···················· |
| L | 8656 | hCoV-19/England/QEUH-9DAE57/2020\|EPI_ISL_580952\|20 | ···················· | ·A·················· | ····················· | ···················· | ···················· | ···················· |
| L | 8657 | hCoV-19/England/QEUH-9DACF3/2020\|EPI_ISL_580953\|20 | ···················· | ···················· | ····················· | ···················· | ···················· | ···················· |

| | | | | | | | |
|---|---|---|---|---|---|---|---|
| L | 8658 hCoV-19/England/QEUH-9DACD5/2020\|EPI_ISL_580954\|20 | ···················· | ···················· | ····················· | ···················· | ···················· | ···················· |
| L | 8659 hCoV-19/England/QEUH-9DC9CD/2020\|EPI_ISL_580955\|20 | ···················· | ···················· | ····················· | ···················· | ···················· | ···················· |
| L | 8660 hCoV-19/England/QEUH-9DC75A/2020\|EPI_ISL_580957\|20 | ···················· | ···················· | ····················· | ···················· | ···················· | ···················· |
| L | 8661 hCoV-19/England/QEUH-9DC7A5/2020\|EPI_ISL_580958\|20 | ···················· | ···················· | ····················· | ···················· | ···················· | ···················· |
| L | 8662 hCoV-19/USA/CA-CZB-10927/2020\|EPI_ISL_583156\|2020- | ···················· | ·A·················· | ····················· | ···················· | ···················· | ···················· |
| L | 8663 hCoV-19/USA/CA-CZB-10928/2020\|EPI_ISL_583157\|2020- | ···················· | ·A·················· | ····················· | ···················· | ···················· | ···················· |
| L | 8664 hCoV-19/USA/CA-CZB-10929/2020\|EPI_ISL_583158\|2020- | ···················· | ·A·················· | ····················· | ···················· | ···················· | ···················· |
| L | 8665 hCoV-19/USA/CA-CZB-10930/2020\|EPI_ISL_583159\|2020- | ···················· | ·A·················· | ····················· | ···················· | ···················· | ···················· |
| L | 8667 hCoV-19/USA/CA-CZB-10933/2020\|EPI_ISL_583162\|2020- | ···················· | ·A·················· | ····················· | ···················· | ···················· | ···················· |
| L | 8668 hCoV-19/USA/CA-CZB-10934/2020\|EPI_ISL_583163\|2020- | ···················· | ···················· | ····················· | ···················· | ···················· | ···················· |
| L | 8670 hCoV-19/USA/CA-CZB-10936/2020\|EPI_ISL_583165\|2020- | ···················· | ·A·················· | ····················· | ···················· | ···················· | ···················· |
| L | 8671 hCoV-19/USA/CA-CZB-10937/2020\|EPI_ISL_583166\|2020- | ···················· | ·A·················· | ····················· | ···················· | ···················· | ···················· |
| L | 8672 hCoV-19/USA/CA-CZB-10938/2020\|EPI_ISL_583167\|2020- | ···················· | ·A·················· | ····················· | ···················· | ···················· | ···················· |
| L | 8673 hCoV-19/USA/CA-CZB-10939/2020\|EPI_ISL_583168\|2020- | ···················· | ·A·················· | ····················· | ···················· | ···················· | ···················· |
| L | 8674 hCoV-19/USA/CA-CZB-10940/2020\|EPI_ISL_583169\|2020- | ···················· | ·A·················· | ····················· | ···················· | ···················· | ···················· |
| L | 8675 hCoV-19/USA/CA-CZB-10941/2020\|EPI_ISL_583170\|2020- | ···················· | ·A·················· | ····················· | ···················· | ···················· | ···················· |
| L | 8676 hCoV-19/USA/CA-CZB-10965/2020\|EPI_ISL_583171\|2020- | ···················· | ·A·················· | ····················· | ···················· | ···················· | ···················· |
| L | 8677 hCoV-19/USA/CA-CZB-10966/2020\|EPI_ISL_583172\|2020- | ···················· | ·A·················· | ····················· | ···················· | ···················· | ···················· |
| L | 8678 hCoV-19/USA/CA-CZB-174/2020\|EPI_ISL_583173\|2020-05 | ···················· | ···················· | ····················· | ···················· | ···················· | ···················· |
| L | 8679 hCoV-19/USA/CA-CZB-176/2020\|EPI_ISL_583174\|2020-05 | ···················· | ···················· | ····················· | ···················· | ···················· | ···················· |
| L | 8680 hCoV-19/USA/CA-CZB-177/2020\|EPI_ISL_583175\|2020-05 | ···················· | ···················· | ····················· | ···················· | ···················· | ···················· |
| L | 8681 hCoV-19/USA/CA-CZB-178/2020\|EPI_ISL_583176\|2020-05 | ···················· | ···················· | ····················· | ···················· | ···················· | ···················· |
| L | 8682 hCoV-19/USA/CA-CZB-180/2020\|EPI_ISL_583177\|2020-05 | ···················· | ···················· | ····················· | ···················· | ···················· | ···················· |
| L | 8683 hCoV-19/USA/CA-CZB-181/2020\|EPI_ISL_583178\|2020-05 | ···················· | ···················· | ····················· | ···················· | ···················· | ···················· |
| L | 8684 hCoV-19/USA/CA-CZB-182/2020\|EPI_ISL_583179\|2020-05 | ···················· | ···················· | ····················· | ···················· | ···················· | ···················· |
| L | 8685 hCoV-19/USA/CA-CZB-183/2020\|EPI_ISL_583180\|2020-05 | ···················· | ···················· | ····················· | ···················· | ···················· | ···················· |
| L | 8686 hCoV-19/USA/CA-CZB-185/2020\|EPI_ISL_583181\|2020-05 | ···················· | ···················· | ····················· | ···················· | ···················· | ···················· |
| L | 8687 hCoV-19/USA/CA-CZB-186/2020\|EPI_ISL_583182\|2020-05 | ···················· | ···················· | ····················· | ···················· | ···················· | ···················· |
| L | 8688 hCoV-19/USA/CA-CZB-187/2020\|EPI_ISL_583183\|2020-05 | ···················· | ···················· | ····················· | ···················· | ···················· | ···················· |
| L | 8689 hCoV-19/USA/CA-CZB-188/2020\|EPI_ISL_583184\|2020-05 | ···················· | ···················· | ····················· | ···················· | ···················· | ···················· |
| L | 8690 hCoV-19/USA/CA-CZB-189/2020\|EPI_ISL_583185\|2020-05 | ···················· | ···················· | ····················· | ···················· | ···················· | ···················· |
| L | 8691 hCoV-19/USA/CA-CZB-190/2020\|EPI_ISL_583186\|2020-05 | ···················· | ···················· | ····················· | ···················· | ···················· | ···················· |
| L | 8692 hCoV-19/USA/CA-CZB-6113/2020\|EPI_ISL_583187\|2020-0 | ···················· | ···················· | ····················· | ···················· | ···················· | ···················· |
| L | 8693 hCoV-19/USA/CA-CZB-6117/2020\|EPI_ISL_583188\|2020-0 | ···················· | ···················· | ·············N······· | ···················· | ···················· | ···················· |
| L | 8694 hCoV-19/USA/CA-CZB-6122/2020\|EPI_ISL_583189\|2020-0 | ···················· | ···················· | ····················· | ···················· | ···················· | ···················· |
| L | 8695 hCoV-19/USA/CA-CZB-10526/2020\|EPI_ISL_583190\|2020- | ···················· | ···················· | ····················· | ···················· | ···················· | ···················· |
| L | 8696 hCoV-19/USA/CA-CZB-10534/2020\|EPI_ISL_583191\|2020- | ···················· | ·A·················· | ····················· | ···················· | ···················· | ···················· |
| L | 8697 hCoV-19/USA/CA-CZB-10542/2020\|EPI_ISL_583192\|2020- | ···················· | ···················· | ····················· | ···················· | ···················· | ···················· |
| L | 8698 hCoV-19/USA/CA-CZB-10558/2020\|EPI_ISL_583193\|2020- | ···················· | ···················· | ····················· | ···················· | ···················· | ···················· |
| L | 8699 hCoV-19/USA/CA-CZB-10566/2020\|EPI_ISL_583194\|2020- | ···················· | ···················· | ····················· | ···················· | ···················· | ···················· |
| L | 8700 hCoV-19/USA/CA-CZB-10582/2020\|EPI_ISL_583195\|2020- | ···················· | ···················· | ····················· | ···················· | ···················· | ···················· |
| L | 8701 hCoV-19/USA/CA-CZB-10590/2020\|EPI_ISL_583196\|2020- | ···················· | ···················· | ····················· | ···················· | ···················· | ···················· |
| L | 8702 hCoV-19/USA/CA-CZB-10598/2020\|EPI_ISL_583197\|2020- | ···················· | ···················· | ····················· | ···················· | ···················· | ···················· |
| L | 8703 hCoV-19/USA/CA-CZB-7028/2020\|EPI_ISL_583198\|2020-0 | ···················· | ·A·················· | ····················· | ···················· | ···················· | ···················· |
| L | 8704 hCoV-19/USA/CA-CZB-193/2020\|EPI_ISL_583199\|2020-08 | ···················· | ···················· | ····················· | ···················· | ···················· | ···················· |
| L | 8705 hCoV-19/USA/MI-QDX-357/2020\|EPI_ISL_571100\|2020-03 | ···················· | ···················· | ····················· | ···················· | ···················· | ···················· |
| L | 8706 hCoV-19/USA/MI-QDX-358/2020\|EPI_ISL_571101\|2020-03 | ···················· | ···················· | ····················· | ···················· | ···················· | ···················· |
| L | 8707 hCoV-19/USA/MI-QDX-359/2020\|EPI_ISL_571102\|2020-03 | ···················· | ···················· | ····················· | ···················· | ···················· | ···················· |
| L | 8708 hCoV-19/USA/MI-QDX-360/2020\|EPI_ISL_571103\|2020-03 | ···················· | ···················· | ····················· | ···················· | ···················· | ···················· |
| L | 8709 hCoV-19/USA/MI-QDX-361/2020\|EPI_ISL_571104\|2020-03 | ···················· | ···················· | ····················· | ···················· | ···················· | ···················· |
| L | 8710 hCoV-19/USA/MI-QDX-362/2020\|EPI_ISL_571105\|2020-03 | ···················· | ···················· | ····················· | ···················· | ···················· | ···················· |
| L | 8711 hCoV-19/USA/MI-QDX-363/2020\|EPI_ISL_571106\|2020-03 | ···T················ | ···················· | ····················· | ···················· | ···················· | ···················· |
| L | 8712 hCoV-19/USA/MI-QDX-364/2020\|EPI_ISL_571107\|2020-03 | ···················· | ···················· | ····················· | ···················· | ···················· | ···················· |
| L | 8713 hCoV-19/USA/MI-QDX-365/2020\|EPI_ISL_571108\|2020-03 | ···················· | ···················· | ····················· | ···················· | ···················· | ···················· |
| L | 8714 hCoV-19/USA/MI-QDX-366/2020\|EPI_ISL_571109\|2020-03 | ···················· | ···················· | ····················· | ···················· | ···················· | ···················· |
| L | 8715 hCoV-19/USA/MI-QDX-367/2020\|EPI_ISL_571110\|2020-03 | ···················· | ···················· | ····················· | ···················· | ···················· | ···················· |
| L | 8716 hCoV-19/USA/MI-QDX-368/2020\|EPI_ISL_571111\|2020-03 | ···················· | ···················· | ····················· | ···················· | ···················· | ···················· |
| L | 8717 hCoV-19/USA/MI-QDX-369/2020\|EPI_ISL_571112\|2020-03 | ···················· | ···················· | ····················· | ···················· | ···················· | ···················· |
| L | 8718 hCoV-19/USA/MI-QDX-370/2020\|EPI_ISL_571113\|2020-03 | ···················· | ···················· | ····················· | ···················· | ···················· | ···················· |
| L | 8719 hCoV-19/USA/MI-QDX-371/2020\|EPI_ISL_571114\|2020-03 | ···················· | ···················· | ····················· | ···················· | ···················· | ···················· |
| L | 8720 hCoV-19/USA/MI-QDX-372/2020\|EPI_ISL_571115\|2020-03 | ···················· | ···················· | ····················· | ···················· | ···················· | ···················· |
| L | 8721 hCoV-19/USA/MI-QDX-373/2020\|EPI_ISL_571116\|2020-03 | ···················· | ···················· | ····················· | ···················· | ···················· | ···················· |
| L | 8722 hCoV-19/USA/MI-QDX-374/2020\|EPI_ISL_571117\|2020-03 | ···················· | ···················· | ····················· | ···················· | ···················· | ···················· |
| L | 8723 hCoV-19/USA/MI-QDX-375/2020\|EPI_ISL_571118\|2020-03 | ···················· | ···················· | ····················· | ···················· | ···················· | ···················· |
| L | 8724 hCoV-19/USA/MI-QDX-376/2020\|EPI_ISL_571119\|2020-03 | ···················· | ···················· | ····················· | ···················· | ···················· | ···················· |
| L | 8725 hCoV-19/USA/MI-QDX-378/2020\|EPI_ISL_571120\|2020-03 | ···················· | ···················· | ····················· | ···················· | ···················· | ···················· |
| L | 8726 hCoV-19/USA/MI-QDX-379/2020\|EPI_ISL_571121\|2020-03 | ···················· | ···················· | ····················· | ···················· | ···················· | ···················· |
| L | 8727 hCoV-19/USA/MI-QDX-380/2020\|EPI_ISL_571122\|2020-03 | ···················· | ···················· | ····················· | ···················· | ···················· | ···················· |
| L | 8728 hCoV-19/USA/MI-QDX-381/2020\|EPI_ISL_571123\|2020-03 | ···················· | ···················· | ····················· | ···················· | ···················· | ···················· |
| L | 8729 hCoV-19/USA/MI-QDX-382/2020\|EPI_ISL_571124\|2020-03 | ···················· | ···················· | ····················· | ···················· | ···················· | ···················· |
| L | 8730 hCoV-19/USA/MI-QDX-383/2020\|EPI_ISL_571125\|2020-03 | ···················· | ···················· | ····················· | ···················· | ···················· | ···················· |
| L | 8731 hCoV-19/USA/MI-QDX-384/2020\|EPI_ISL_571126\|2020-03 | ···················· | ···················· | ····················· | ···················· | ···················· | ···················· |
| L | 8732 hCoV-19/USA/MI-QDX-385/2020\|EPI_ISL_571127\|2020-03 | ···················· | ···················· | ····················· | ···················· | ···················· | ···················· |
| L | 8733 hCoV-19/USA/MI-QDX-386/2020\|EPI_ISL_571128\|2020-03 | ···················· | ···················· | ····················· | ···················· | ···················· | ···················· |
| L | 8734 hCoV-19/USA/MI-QDX-388/2020\|EPI_ISL_571129\|2020-03 | ···················· | ···················· | ····················· | ···················· | ···················· | ···················· |
| L | 8735 hCoV-19/USA/MI-QDX-389/2020\|EPI_ISL_571130\|2020-03 | ···················· | ···················· | ····················· | ···················· | ···················· | ···················· |
| L | 8736 hCoV-19/USA/MI-QDX-390/2020\|EPI_ISL_571131\|2020-03 | ···················· | ···················· | ····················· | ···················· | ···················· | ···················· |
| L | 8737 hCoV-19/USA/MI-QDX-391/2020\|EPI_ISL_571132\|2020-03 | ···················· | ···················· | ····················· | ···················· | ···················· | ···················· |
| L | 8738 hCoV-19/USA/MI-QDX-392/2020\|EPI_ISL_571133\|2020-03 | ···················· | ···················· | ····················· | ···················· | ···················· | ···················· |
| L | 8739 hCoV-19/USA/MI-QDX-393/2020\|EPI_ISL_571134\|2020-03 | ···················· | ···················· | ····················· | ···················· | ···················· | ···················· |

| | | | | | | | |
|---|---|---|---|---|---|---|---|
| L | 8740 hCoV-19/USA/MI-QDX-395/2020\|EPI_ISL_571135\|2020-03 | ···················· | ···················· | ···················· | ···················· | ···················· | ···················· |
| L | 8741 hCoV-19/USA/MI-QDX-397/2020\|EPI_ISL_571136\|2020-03 | ···················· | ···················· | ···················· | ···················· | ···················· | ···················· |
| L | 8742 hCoV-19/USA/MI-QDX-398/2020\|EPI_ISL_571137\|2020-03 | ···················· | ···················· | ···················· | ···················· | ···················· | ···················· |
| L | 8743 hCoV-19/USA/MI-QDX-399/2020\|EPI_ISL_571138\|2020-03 | ···················· | ···················· | ···················· | ···················· | ···················· | ···················· |
| L | 8744 hCoV-19/USA/MI-QDX-400/2020\|EPI_ISL_571139\|2020-03 | ···················· | ···················· | ···················· | ···················· | ···················· | ···················· |
| L | 8745 hCoV-19/USA/MI-QDX-401/2020\|EPI_ISL_571140\|2020-03 | ···················· | ···················· | ···················· | ···················· | ···················· | ···················· |
| L | 8746 hCoV-19/USA/MI-QDX-402/2020\|EPI_ISL_571141\|2020-03 | ···················· | ···················· | ···················· | ···················· | ···················· | ···················· |
| L | 8747 hCoV-19/USA/MI-QDX-403/2020\|EPI_ISL_571142\|2020-03 | ···················· | ···················· | ···················· | ···················· | ···················· | ···················· |
| L | 8748 hCoV-19/USA/MI-QDX-404/2020\|EPI_ISL_571143\|2020-03 | ···················· | ···················· | ···················· | ···················· | ···················· | ···················· |
| L | 8749 hCoV-19/USA/MI-QDX-405/2020\|EPI_ISL_571144\|2020-03 | ···················· | ···················· | ···················· | ···················· | ···················· | ···················· |
| L | 8750 hCoV-19/USA/MI-QDX-406/2020\|EPI_ISL_571145\|2020-03 | ···················· | ···················· | ···················· | ···················· | ···················· | ···················· |
| L | 8751 hCoV-19/USA/MI-QDX-407/2020\|EPI_ISL_571146\|2020-03 | ···················· | ···················· | ···················· | ···················· | ···················· | ···················· |
| L | 8752 hCoV-19/USA/MI-QDX-408/2020\|EPI_ISL_571147\|2020-03 | ···················· | ···················· | ···················· | ···················· | ···················· | ···················· |
| L | 8753 hCoV-19/USA/MI-QDX-409/2020\|EPI_ISL_571148\|2020-03 | ···················· | ···················· | ···················· | ···················· | ···················· | ···················· |
| L | 8754 hCoV-19/USA/TX-QDX-270/2020\|EPI_ISL_571149\|2020-03 | ···················· | ···················· | ···················· | ···················· | ···················· | ···················· |
| L | 8755 hCoV-19/USA/TX-QDX-271/2020\|EPI_ISL_571150\|2020-03 | ···················· | ···················· | ···················· | ···················· | ···················· | ···················· |
| L | 8756 hCoV-19/USA/TX-QDX-355/2020\|EPI_ISL_571151\|2020-03 | ···················· | ···················· | ···················· | ···················· | ···················· | ···················· |
| L | 8757 hCoV-19/USA/WA-QDX-276/2020\|EPI_ISL_571152\|2020-03 | ···················· | ···················· | ···················· | ···················· | ···················· | ···················· |
| L | 8758 hCoV-19/USA/WA-QDX-288/2020\|EPI_ISL_571153\|2020-03 | ···················· | ···················· | ···················· | ···················· | ···················· | ···················· |
| L | 8759 hCoV-19/USA/WA-QDX-290/2020\|EPI_ISL_571154\|2020-03 | ···················· | ···················· | ···················· | ···················· | ···················· | ···················· |
| L | 8760 hCoV-19/USA/AL-QDX-470/2020\|EPI_ISL_571155\|2020-03 | ···················· | ···················· | ···················· | ···················· | ···················· | ···················· |
| L | 8761 hCoV-19/USA/CA-QDX-423/2020\|EPI_ISL_571156\|2020-03 | ···················· | ···················· | ···················· | ···················· | ···················· | ···················· |
| L | 8762 hCoV-19/USA/CA-QDX-425/2020\|EPI_ISL_571157\|2020-03 | ···················· | ···················· | ···················· | ···················· | ···················· | ···················· |
| L | 8763 hCoV-19/USA/CA-QDX-433/2020\|EPI_ISL_571158\|2020-03 | ···················· | ···················· | ···················· | ···················· | ···················· | ···················· |
| L | 8764 hCoV-19/USA/CA-QDX-434/2020\|EPI_ISL_571159\|2020-03 | ···················· | ···················· | ···················· | ···················· | ···················· | ···················· |
| L | 8765 hCoV-19/USA/CA-QDX-435/2020\|EPI_ISL_571160\|2020-03 | ···················· | ···················· | ···················· | ···················· | ···················· | ···················· |
| L | 8766 hCoV-19/USA/CA-QDX-436/2020\|EPI_ISL_571161\|2020-03 | ···················· | ···················· | ···················· | ···················· | ···················· | ···················· |
| L | 8767 hCoV-19/USA/CA-QDX-437/2020\|EPI_ISL_571162\|2020-03 | ···················· | ···················· | ···················· | ···················· | ···················· | ···················· |
| L | 8768 hCoV-19/USA/CA-QDX-441/2020\|EPI_ISL_571163\|2020-03 | ···················· | ···················· | ···················· | ···················· | ···················· | ···················· |
| L | 8769 hCoV-19/USA/CA-QDX-442/2020\|EPI_ISL_571164\|2020-03 | ···················· | ···················· | ···················· | ···················· | ···················· | ···················· |
| L | 8770 hCoV-19/USA/CA-QDX-443/2020\|EPI_ISL_571165\|2020-03 | ···················· | ···················· | ···················· | ···················· | ···················· | ···················· |
| L | 8771 hCoV-19/USA/CA-QDX-444/2020\|EPI_ISL_571166\|2020-03 | ···················· | ···················· | ···················· | ···················· | ···················· | ···················· |
| L | 8772 hCoV-19/USA/CA-QDX-445/2020\|EPI_ISL_571167\|2020-03 | ···················· | ···················· | ···················· | ···················· | ···················· | ···················· |
| L | 8773 hCoV-19/USA/CA-QDX-446/2020\|EPI_ISL_571168\|2020-03 | ···················· | ···················· | ···················· | ···················· | ···················· | ···················· |
| L | 8774 hCoV-19/USA/CA-QDX-447/2020\|EPI_ISL_571169\|2020-03 | ···················· | ···················· | ···················· | ···················· | ···················· | ···················· |
| L | 8775 hCoV-19/USA/CA-QDX-448/2020\|EPI_ISL_571170\|2020-03 | ···················· | ···················· | ···················· | ···················· | ···················· | ···················· |
| L | 8776 hCoV-19/USA/CA-QDX-449/2020\|EPI_ISL_571171\|2020-03 | ···················· | ···················· | ···················· | ···················· | ···················· | ···················· |
| L | 8777 hCoV-19/USA/CA-QDX-450/2020\|EPI_ISL_571172\|2020-03 | ···················· | ···················· | ···················· | ···················· | ···················· | ···················· |
| L | 8778 hCoV-19/USA/CA-QDX-452/2020\|EPI_ISL_571174\|2020-03 | ···················· | ···················· | ···················· | ···················· | ···················· | ···················· |
| L | 8779 hCoV-19/USA/CA-QDX-453/2020\|EPI_ISL_571175\|2020-03 | ···················· | ···················· | ···················· | ···················· | ···················· | ···················· |
| L | 8780 hCoV-19/USA/CA-QDX-513/2020\|EPI_ISL_571176\|2020-03 | ···················· | ···················· | ···················· | ···················· | ···················· | ···················· |
| L | 8781 hCoV-19/England/QEUH-9BA752/2020\|EPI_ISL_573600\|20 | ···················· | ···················· | ···················· | ···················· | ···················· | ···················· |
| L | 8782 hCoV-19/England/QEUH-9BA770/2020\|EPI_ISL_573601\|20 | ···················· | ···················· | ···················· | ···················· | ···················· | ···················· |
| L | 8783 hCoV-19/England/QEUH-9BA79E/2020\|EPI_ISL_573602\|20 | ···················· | ···················· | ···················· | ···················· | ···················· | ···················· |
| L | 8784 hCoV-19/England/QEUH-9BA7AD/2020\|EPI_ISL_573603\|20 | ···················· | ···················· | ···················· | ···················· | ···················· | ···················· |
| L | 8785 hCoV-19/England/QEUH-9BA822/2020\|EPI_ISL_573604\|20 | ···················· | ···················· | ···················· | ···················· | ···················· | ···················· |
| L | 8786 hCoV-19/England/QEUH-9BA831/2020\|EPI_ISL_573605\|20 | ···················· | ·A·················· | ···················· | ···················· | ···················· | ···················· |
| L | 8787 hCoV-19/England/QEUH-9BA840/2020\|EPI_ISL_573606\|20 | ···················· | ···················· | ···················· | ···················· | ···················· | ···················· |
| L | 8788 hCoV-19/England/QEUH-9BA89B/2020\|EPI_ISL_573607\|20 | ···················· | ···················· | ···················· | ···················· | ···················· | ···················· |
| L | 8789 hCoV-19/England/QEUH-9BA8AA/2020\|EPI_ISL_573608\|20 | ···················· | ···················· | ···················· | ···················· | ······T············· | ···················· |
| L | 8790 hCoV-19/England/QEUH-9BA8B9/2020\|EPI_ISL_573609\|20 | ···················· | ···················· | ···················· | ···················· | ···················· | ···················· |
| L | 8791 hCoV-19/England/SHEF-D1581/2020\|EPI_ISL_573751\|202 | ···················· | ···················· | ···················· | ···················· | ···················· | ···················· |
| L | 8792 hCoV-19/England/SHEF-D1EC2/2020\|EPI_ISL_573752\|202 | ···················· | ···················· | ···················· | ···················· | ···················· | ···················· |
| L | 8793 hCoV-19/England/SHEF-D2C9A/2020\|EPI_ISL_573753\|202 | ···················· | ···················· | ···················· | ···················· | ···················· | ···················· |
| L | 8794 hCoV-19/England/SHEF-D2CB8/2020\|EPI_ISL_573754\|202 | ···················· | ·A·················· | ···················· | ···················· | ···················· | ···················· |
| L | 8795 hCoV-19/England/SHEF-D30D9/2020\|EPI_ISL_573755\|202 | ···················· | ···················· | ···················· | ···················· | ···················· | ···················· |
| L | 8796 hCoV-19/England/SHEF-D3112/2020\|EPI_ISL_573756\|202 | ···················· | ·A·················· | ···················· | ···················· | ···················· | ···A················ |
| L | 8797 hCoV-19/England/SHEF-D316D/2020\|EPI_ISL_573757\|202 | ···················· | ···················· | ···················· | ···················· | ···················· | ···················· |
| L | 8798 hCoV-19/England/SHEF-D3367/2020\|EPI_ISL_573758\|202 | ···················· | ···················· | ···················· | ···················· | ···················· | ···················· |
| L | 8799 hCoV-19/Northern Ireland/CAMC-9BFDB5/2020\|EPI_ISL_ | ···················· | ···················· | ···················· | ···················· | ···················· | ···················· |
| L | 8800 hCoV-19/Northern Ireland/MILK-9C71BE/2020\|EPI_ISL_ | ···················· | ···················· | ···················· | ···················· | ······T············· | ···················· |
| L | 8801 hCoV-19/Northern Ireland/MILK-9C71CD/2020\|EPI_ISL_ | ···················· | ···················· | ···················· | ···················· | ···················· | ···················· |
| L | 8802 hCoV-19/Northern Ireland/QEUH-9B3C5A/2020\|EPI_ISL_ | ···················· | ···················· | ···················· | ···················· | ···················· | ···················· |
| L | 8803 hCoV-19/Northern Ireland/QEUH-9B3DB1/2020\|EPI_ISL_ | ···················· | ···················· | ···················· | ···················· | ···················· | ···················· |
| L | 8804 hCoV-19/Northern Ireland/QEUH-9D1BD1/2020\|EPI_ISL_ | ···················· | ···················· | ···················· | ···················· | ···················· | ···················· |
| L | 8805 hCoV-19/Scotland/CVR4112/2020\|EPI_ISL_573765\|2020- | ···················· | ···················· | ···················· | ···················· | ···················· | ···················· |
| L | 8806 hCoV-19/Scotland/CVR4116/2020\|EPI_ISL_573766\|2020- | ···················· | ···················· | ···················· | ···················· | ···················· | ···················· |
| L | 8808 hCoV-19/Scotland/CVR4126/2020\|EPI_ISL_573768\|2020- | ···················· | ···················· | ···················· | ···················· | ···················· | ···················· |
| L | 8809 hCoV-19/Scotland/CVR4296/2020\|EPI_ISL_573769\|2020- | ···················· | ···················· | ···················· | ···················· | ···················· | ···················· |
| L | 8810 hCoV-19/Scotland/CVR4309/2020\|EPI_ISL_573770\|2020- | ···················· | ···················· | ···················· | ···················· | ···················· | ···················· |
| L | 8812 hCoV-19/Scotland/CVR4322/2020\|EPI_ISL_573773\|2020- | ···················· | ···················· | ···················· | ···················· | ···················· | ···················· |
| L | 8813 hCoV-19/Scotland/CVR4323/2020\|EPI_ISL_573774\|2020- | ···················· | ···················· | ···················· | ···················· | ···················· | ···················· |
| L | 8814 hCoV-19/Scotland/CVR4326/2020\|EPI_ISL_573775\|2020- | ···················· | ···················· | ···················· | ···················· | ···················· | NN·················· |
| L | 8815 hCoV-19/Scotland/CVR4329/2020\|EPI_ISL_573776\|2020- | ···················· | ···················· | ···················· | ···················· | ···················· | ···················· |
| L | 8816 hCoV-19/Scotland/CVR4331/2020\|EPI_ISL_573777\|2020- | ···················· | ···················· | ···················· | ···················· | ···················· | ···················· |
| L | 8818 hCoV-19/Scotland/CVR4338/2020\|EPI_ISL_573779\|2020- | ···················· | ···················· | ···················· | ···················· | ···················· | ···················· |
| L | 8819 hCoV-19/Scotland/CVR4339/2020\|EPI_ISL_573780\|2020- | ···················· | ···················· | ···················· | ···················· | ···················· | ···················· |
| L | 8820 hCoV-19/Scotland/CVR4343/2020\|EPI_ISL_573781\|2020- | ···················· | ···················· | ···················· | ···················· | ···················· | ···················· |
| L | 8821 hCoV-19/Scotland/CVR4344/2020\|EPI_ISL_573782\|2020- | ···················· | ···················· | ···················· | ···················· | ···················· | ···················· |
| L | 8822 hCoV-19/Scotland/CVR4345/2020\|EPI_ISL_573783\|2020- | ···················· | ···················· | ···················· | ···················· | ··············N····· | NN·················· |

| | | | | | | | | | |
|---|---|---|---|---|---|---|---|---|---|
| L | 8823 | hCoV-19/Scotland/CVR4352/2020\|EPI_ISL_573784\|2020- | ···················· | ···················· | ···················· | ···················· | ···················· | NN·················· | |
| L | 8824 | hCoV-19/Scotland/CVR4356/2020\|EPI_ISL_573785\|2020- | ···················· | ···················· | ···················· | ···················· | ···················· | ··N················· | |
| L | 8825 | hCoV-19/Scotland/CVR4361/2020\|EPI_ISL_573786\|2020- | ···················· | ···················· | ···················· | ···················· | ···················· | ··N·······N········· | |
| L | 8826 | hCoV-19/Scotland/CVR4364/2020\|EPI_ISL_573787\|2020- | ···················· | ···················· | ···················· | ···················· | ···················· | ···················· | |
| L | 8827 | hCoV-19/Scotland/CVR4365/2020\|EPI_ISL_573788\|2020- | ···················· | ···················· | ···················· | ···················· | ···················· | ···················· | |
| L | 8828 | hCoV-19/Scotland/CVR4367/2020\|EPI_ISL_573789\|2020- | ···················· | ···················· | ···················· | ···················· | ···················· | ···················· | |
| L | 8829 | hCoV-19/Scotland/CVR4368/2020\|EPI_ISL_573790\|2020- | ···················· | ···················· | ···················· | ···················· | ···················· | ···················· | |
| L | 8830 | hCoV-19/Scotland/CVR4369/2020\|EPI_ISL_573791\|2020- | ···················· | ···················· | ···················· | ···················· | ···················· | ···················· | |
| L | 8831 | hCoV-19/Scotland/CVR4373/2020\|EPI_ISL_573792\|2020- | ···················· | ···················· | ···················· | ···················· | ···················· | ···················· | |
| L | 8832 | hCoV-19/Scotland/CVR4374/2020\|EPI_ISL_573793\|2020- | ···················· | ···················· | ···················· | ···················· | ···················· | ···················· | |
| L | 8833 | hCoV-19/Scotland/EDB7772/2020\|EPI_ISL_573794\|2020- | ···················· | ···················· | ···················· | ···················· | ···················· | ···················· | |
| L | 8835 | hCoV-19/Scotland/EDB7820/2020\|EPI_ISL_573796\|2020- | ···················· | ·A·················· | ···················· | ···················· | ···················· | ···················· | |
| L | 8836 | hCoV-19/Scotland/EDB7843/2020\|EPI_ISL_573797\|2020- | ···················· | ···················· | ···················· | ···················· | ···················· | ···················· | |
| L | 8837 | hCoV-19/Scotland/EDB7846/2020\|EPI_ISL_573798\|2020- | ···················· | ···················· | ···················· | ···················· | ···················· | ···················· | |
| L | 8839 | hCoV-19/Switzerland/BL-ETHZ-100356/2020\|EPI_ISL_57 | ···················· | ···················· | ···················· | ···················· | ···················· | ···················· | |
| L | 8840 | hCoV-19/Switzerland/BL-ETHZ-100363/2020\|EPI_ISL_57 | ···················· | ···················· | ···················· | ···················· | ···················· | ···················· | |
| L | 8841 | hCoV-19/Switzerland/BL-ETHZ-100364/2020\|EPI_ISL_57 | ···················· | ···················· | ···················· | ···················· | ···················· | ···················· | |
| L | 8842 | hCoV-19/Switzerland/BS-ETHZ-100158/2020\|EPI_ISL_57 | ···················· | ···················· | ···················· | ···················· | ···················· | ···················· | |
| L | 8843 | hCoV-19/Switzerland/BS-ETHZ-100199/2020\|EPI_ISL_57 | ···················· | ···················· | ···················· | ···················· | ···················· | ···················· | |
| L | 8844 | hCoV-19/Switzerland/BS-ETHZ-100201/2020\|EPI_ISL_57 | ···················· | ···················· | ···················· | ···················· | ···················· | ···················· | |
| L | 8845 | hCoV-19/Switzerland/BS-ETHZ-100203/2020\|EPI_ISL_57 | ···················· | ···················· | ···················· | ···················· | ···················· | ····M··············· | |
| L | 8846 | hCoV-19/Switzerland/BS-ETHZ-100212/2020\|EPI_ISL_57 | ···················· | ···················· | ···················· | ···················· | ···················· | ···················· | |
| L | 8847 | hCoV-19/Switzerland/BS-ETHZ-100241/2020\|EPI_ISL_57 | ···················· | ···················· | ···················· | ···················· | ···················· | ···················· | |
| L | 8848 | hCoV-19/Switzerland/BS-ETHZ-100383/2020\|EPI_ISL_57 | ···················· | ···················· | ···················· | ···················· | ···················· | ···················· | |
| L | 8849 | hCoV-19/Switzerland/BE-ETHZ-100160/2020\|EPI_ISL_57 | ···················· | ···················· | ···················· | ···················· | ···················· | ···················· | |
| L | 8850 | hCoV-19/Switzerland/BE-ETHZ-100161/2020\|EPI_ISL_57 | ···················· | ···················· | ···················· | ···················· | ···················· | ···················· | |
| L | 8851 | hCoV-19/Switzerland/BE-ETHZ-100197/2020\|EPI_ISL_57 | ···················· | ···················· | ···················· | ···················· | ···················· | ···················· | |
| L | 8852 | hCoV-19/Switzerland/BE-ETHZ-100213/2020\|EPI_ISL_57 | ···················· | ···················· | ···················· | ···················· | ···················· | ···················· | |
| L | 8853 | hCoV-19/Switzerland/BE-ETHZ-100216/2020\|EPI_ISL_57 | ···················· | ···················· | ···················· | ···················· | ···················· | ···················· | |
| L | 8854 | hCoV-19/Switzerland/BE-ETHZ-100218/2020\|EPI_ISL_57 | ···················· | ···················· | ···················· | ···················· | ···················· | ···················· | |
| L | 8855 | hCoV-19/Switzerland/BE-ETHZ-100219/2020\|EPI_ISL_57 | ···················· | ···················· | ···················· | ···················· | ···················· | ···················· | |
| L | 8856 | hCoV-19/Switzerland/BE-ETHZ-100237/2020\|EPI_ISL_57 | ···················· | ···················· | ···················· | ···················· | ···················· | ···················· | |
| L | 8857 | hCoV-19/Switzerland/BE-ETHZ-100341/2020\|EPI_ISL_57 | ···················· | ···················· | ···················· | ···················· | ···················· | ···················· | |
| L | 8858 | hCoV-19/Switzerland/BE-ETHZ-100349/2020\|EPI_ISL_57 | ···················· | ···················· | ···················· | ···················· | ···················· | ···················· | |
| L | 8859 | hCoV-19/Switzerland/BE-ETHZ-100369/2020\|EPI_ISL_57 | ···················· | ···················· | ···················· | ···················· | ···················· | ···················· | |
| L | 8860 | hCoV-19/Switzerland/BE-ETHZ-100372/2020\|EPI_ISL_57 | ···················· | ···················· | ···················· | ···················· | ···················· | ···················· | |
| L | 8861 | hCoV-19/Switzerland/BE-ETHZ-100376/2020\|EPI_ISL_57 | ···················· | ···················· | ···················· | ···················· | ···················· | ···················· | |
| L | 8862 | hCoV-19/Switzerland/BE-ETHZ-100377/2020\|EPI_ISL_57 | ···················· | ···················· | ···················· | ···················· | ···················· | ···················· | |
| L | 8863 | hCoV-19/Switzerland/BE-ETHZ-100378/2020\|EPI_ISL_57 | ···················· | ···················· | ···················· | ···················· | ···················· | ···················· | |
| L | 8864 | hCoV-19/Switzerland/BE-ETHZ-100382/2020\|EPI_ISL_57 | ···················· | ···················· | ···················· | ···················· | ···················· | ···················· | |
| L | 8865 | hCoV-19/Switzerland/GE-ETHZ-100166/2020\|EPI_ISL_57 | ···················· | ···················A | ···················· | ···················· | ···················· | ···················· | 1 |
| L | 8866 | hCoV-19/Switzerland/GE-ETHZ-100183/2020\|EPI_ISL_57 | ···················· | ···················· | ···················· | ···················· | ···················· | ···················· | |
| L | 8867 | hCoV-19/Switzerland/GE-ETHZ-100188/2020\|EPI_ISL_57 | ···················· | ···················· | ···················· | ···················· | ···················· | ···················· | |
| L | 8868 | hCoV-19/Switzerland/GE-ETHZ-100189/2020\|EPI_ISL_57 | ···················· | ·A·················· | ···················· | ···················· | ···················· | ···················· | |
| L | 8869 | hCoV-19/Switzerland/GE-ETHZ-100309/2020\|EPI_ISL_57 | ···················· | ···················· | ···················· | ···················· | ···················· | ···················· | |
| L | 8870 | hCoV-19/Switzerland/GE-ETHZ-100318/2020\|EPI_ISL_57 | ···················· | ···················· | ···················· | ···················· | ···················· | ···················· | |
| L | 8871 | hCoV-19/Switzerland/GE-ETHZ-100336/2020\|EPI_ISL_57 | ···················· | ···················· | ···················· | ···················· | ···················· | ···················· | |
| L | 8872 | hCoV-19/Switzerland/GE-ETHZ-100358/2020\|EPI_ISL_57 | ···················· | ···················· | ···················· | ···················· | ···················· | ···················· | |
| L | 8873 | hCoV-19/England/NOTT-11432D/2020\|EPI_ISL_577160\|20 | ···················· | ···················· | ···················· | ···················· | ···················· | ···················· | |
| L | 8874 | hCoV-19/England/NOTT-1143D2/2020\|EPI_ISL_577161\|20 | ···················· | ·A·················· | ···················· | ···················· | ···················· | ··A················· | |
| L | 8875 | hCoV-19/England/NOTT-1143F0/2020\|EPI_ISL_577162\|20 | ···················· | ·A·················· | ···················· | ···················· | ···················· | ··A················· | |
| L | 8876 | hCoV-19/England/NOTT-114439/2020\|EPI_ISL_577163\|20 | ···················· | ···················· | ···················· | ···················· | ···················· | ···················· | |
| L | 8877 | hCoV-19/England/NOTT-114475/2020\|EPI_ISL_577165\|20 | ···················· | ···················· | ···················· | ···················· | ···················· | ···················· | |
| L | 8878 | hCoV-19/England/NOTT-114484/2020\|EPI_ISL_577166\|20 | ···················· | ···················· | ···················· | ···················· | ···················· | ···················· | |
| L | 8879 | hCoV-19/England/NOTT-114493/2020\|EPI_ISL_577167\|20 | ···················· | ···················· | ···················· | ···················· | ···················· | ···················· | |
| L | 8880 | hCoV-19/England/NOTT-1144A2/2020\|EPI_ISL_577168\|20 | ···················· | ···················· | ···················· | ···················· | ···················· | ···················· | |
| L | 8881 | hCoV-19/England/NOTT-1144EE/2020\|EPI_ISL_577169\|20 | ···················· | ···················· | ···················· | ···················· | ···················· | ···················· | |
| L | 8882 | hCoV-19/England/NOTT-1144FD/2020\|EPI_ISL_577170\|20 | ···················· | ···G················ | ···················· | ···················· | ···················· | ···················· | |
| L | 8883 | hCoV-19/England/NOTT-114518/2020\|EPI_ISL_577171\|20 | ···················· | ···················· | ···················· | ···················· | ···················· | ···················· | |
| L | 8884 | hCoV-19/England/NOTT-114536/2020\|EPI_ISL_577172\|20 | ···················· | ···················· | ···················· | ···················· | ···················· | ···················· | |
| L | 8885 | hCoV-19/England/NOTT-114554/2020\|EPI_ISL_577173\|20 | ···················· | ···················· | ···················· | ···················· | ···················· | ···················· | |
| L | 8886 | hCoV-19/England/NOTT-114563/2020\|EPI_ISL_577174\|20 | ···················· | ···················· | ···················· | ···················· | ···················· | ···················· | |
| L | 8887 | hCoV-19/England/NOTT-1145AF/2020\|EPI_ISL_577176\|20 | ···················· | ···················· | ···················· | ···················· | ···················· | ···················· | |
| L | 8888 | hCoV-19/England/NOTT-1145CD/2020\|EPI_ISL_577178\|20 | ···················· | ···················· | ···················· | ···················· | ···················· | ···················· | |
| L | 8889 | hCoV-19/England/NOTT-1145DC/2020\|EPI_ISL_577179\|20 | ···················· | ···················· | ···················· | ···················· | ···················· | ···················· | |
| L | 8890 | hCoV-19/England/NOTT-114615/2020\|EPI_ISL_577180\|20 | ···················· | ···················· | ···················· | ···················· | ···················· | ···················· | |
| L | 8891 | hCoV-19/England/NOTT-114624/2020\|EPI_ISL_577181\|20 | ···················· | ···················· | ···················· | ···················· | ···················· | ···················· | |
| L | 8892 | hCoV-19/England/NOTT-114633/2020\|EPI_ISL_577182\|20 | ···················· | ···················· | ···················· | ···················· | ···················· | ···················· | |
| L | 8893 | hCoV-19/England/NOTT-114642/2020\|EPI_ISL_577183\|20 | ···················· | ···················· | ···················· | ···················· | ···················· | ···················· | |
| L | 8894 | hCoV-19/England/NOTT-114660/2020\|EPI_ISL_577184\|20 | ···················· | ···················· | ···················· | ···················· | ·····T·············· | ···················· | |
| L | 8895 | hCoV-19/England/NOTT-11467F/2020\|EPI_ISL_577185\|20 | ···················· | ···················· | ···················· | ···················· | ···················· | ···················· | |
| L | 8896 | hCoV-19/England/NOTT-11468E/2020\|EPI_ISL_577186\|20 | ···················· | ···················· | ···················· | ···················· | ···················· | ···················· | |
| L | 8897 | hCoV-19/England/NOTT-11469D/2020\|EPI_ISL_577187\|20 | ···················· | ·A·················· | ···················· | ···················· | ···················· | ··A················· | |
| L | 8898 | hCoV-19/England/NOTT-114730/2020\|EPI_ISL_577192\|20 | ···················· | ···················· | ···················· | ···················· | ···················· | ···················· | |
| L | 8903 | hCoV-19/USA/WI-WSLH-200313/2020\|EPI_ISL_578494\|202 | ···················· | ···················· | ···················· | ···················· | ···················· | ···················· | |
| L | 8908 | hCoV-19/USA/WI-WSLH-200318/2020\|EPI_ISL_578499\|202 | ···················· | ···················· | ···················· | ···················· | ···················· | ···················· | |
| L | 8909 | hCoV-19/USA/CA-CZB-10856/2020\|EPI_ISL_583100\|2020- | ···················· | ···················· | ···················· | ···················· | ···················· | ···················· | |
| L | 8910 | hCoV-19/USA/CA-CZB-10858/2020\|EPI_ISL_583101\|2020- | ···················· | ···················· | ···················· | ···················· | ···················· | ···················· | |
| L | 8911 | hCoV-19/USA/CA-CZB-10860/2020\|EPI_ISL_583102\|2020- | ···················· | ·A·················· | ···················· | ···················· | ···················· | ···················· | |
| L | 8913 | hCoV-19/USA/CA-CZB-10863/2020\|EPI_ISL_583105\|2020- | ···················· | ·A·················· | ···················· | ···················· | ···················· | ···················· | |

| | | | | | | | | | |
|---|---|---|---|---|---|---|---|---|---|
| L | 8914 | hCoV-19/USA/CA-CZB-10865/2020\|EPI_ISL_583106\|2020- | ···················· | ·A·················· | ···················· | ···················· | ···················· | ···················· | |
| L | 8915 | hCoV-19/USA/CA-CZB-10866/2020\|EPI_ISL_583107\|2020- | ···················· | ·A·················· | ···················· | ···················· | ···················· | ···················· | |
| L | 8916 | hCoV-19/USA/CA-CZB-10867/2020\|EPI_ISL_583108\|2020- | ···················· | ·A·················· | ···················· | ···················· | ···················· | ···················· | |
| L | 8917 | hCoV-19/USA/CA-CZB-10868/2020\|EPI_ISL_583109\|2020- | ···················· | ·A·················· | ···················· | ···················· | ···················· | ···················· | |
| L | 8920 | hCoV-19/USA/CA-CZB-10872/2020\|EPI_ISL_583112\|2020- | ···················· | ···················· | ···················· | ···················· | ···················· | ···················· | |
| L | 8921 | hCoV-19/USA/CA-CZB-10873/2020\|EPI_ISL_583113\|2020- | ···················· | ···················· | ···················· | ···················· | ···················· | ···················· | |
| L | 8922 | hCoV-19/USA/CA-CZB-10874/2020\|EPI_ISL_583114\|2020- | ···················· | ···················· | ···················· | ···················· | ···················· | ···················· | |
| L | 8923 | hCoV-19/USA/CA-CZB-10876/2020\|EPI_ISL_583115\|2020- | ···················· | ···················· | ···················· | ···················· | ···················· | ···················· | |
| L | 8924 | hCoV-19/USA/CA-CZB-10878/2020\|EPI_ISL_583116\|2020- | ···················· | ···················· | ···················· | ···················· | ···················· | ···················· | |
| L | 8926 | hCoV-19/USA/CA-CZB-10880/2020\|EPI_ISL_583118\|2020- | ···················· | ·A·················· | ···················· | ···················· | ··N················· | ···················· | |
| L | 8927 | hCoV-19/USA/CA-CZB-10881/2020\|EPI_ISL_583119\|2020- | ···················· | ·A·················· | ···················· | ···················· | ···················· | ···················· | |
| L | 8928 | hCoV-19/USA/CA-CZB-10885/2020\|EPI_ISL_583120\|2020- | ···················· | ·A·················· | ···················· | ···················· | ···················· | ···················· | |
| L | 8929 | hCoV-19/USA/CA-CZB-10887/2020\|EPI_ISL_583121\|2020- | ···················· | ···················· | ···················· | ···················· | ···················· | ···················· | |
| L | 8930 | hCoV-19/USA/CA-CZB-10889/2020\|EPI_ISL_583122\|2020- | ···················· | ···················· | ···················· | ···················· | ···················· | ···················· | |
| L | 8931 | hCoV-19/USA/CA-CZB-10891/2020\|EPI_ISL_583123\|2020- | ···················· | ···················· | ···················· | ···················· | ···················· | ···················· | |
| L | 8932 | hCoV-19/USA/CA-CZB-10892/2020\|EPI_ISL_583124\|2020- | ···················· | ···················· | ···················· | ···················· | ···················· | ···················· | |
| L | 8933 | hCoV-19/USA/CA-CZB-10895/2020\|EPI_ISL_583125\|2020- | ···················· | ···················· | ···················· | ···················· | ···················· | ···················· | |
| L | 8934 | hCoV-19/USA/CA-CZB-10896/2020\|EPI_ISL_583126\|2020- | ···················· | ···················· | ···················· | ···················· | ···················· | ···················· | |
| L | 8935 | hCoV-19/USA/CA-CZB-10897/2020\|EPI_ISL_583127\|2020- | ···················· | ·A·················· | ···················· | ···················· | ···················· | ···················· | |
| L | 8936 | hCoV-19/USA/CA-CZB-10898/2020\|EPI_ISL_583128\|2020- | ···················· | ·A·················· | ···················· | ···················· | ···················· | ···················· | |
| L | 8937 | hCoV-19/USA/CA-CZB-10899/2020\|EPI_ISL_583129\|2020- | ···················· | ·A·················· | ···················· | ···················· | ···················· | ···················· | |
| L | 8938 | hCoV-19/USA/CA-CZB-10902/2020\|EPI_ISL_583131\|2020- | ···················· | ···················· | ···················· | ···················· | ···················· | ···················· | |
| L | 8939 | hCoV-19/USA/CA-CZB-10903/2020\|EPI_ISL_583132\|2020- | ···················· | ·A·················· | ···················· | ···················· | ···················· | ···················· | |
| L | 8940 | hCoV-19/USA/CA-CZB-10904/2020\|EPI_ISL_583133\|2020- | ···················· | ···················· | ···················· | ···················· | ···················· | ···················· | |
| L | 8941 | hCoV-19/USA/CA-CZB-10905/2020\|EPI_ISL_583134\|2020- | ···················· | ·A·················· | ···················· | ···················· | ···················· | ···················· | |
| L | 8942 | hCoV-19/USA/CA-CZB-10906/2020\|EPI_ISL_583135\|2020- | ···················· | ·A·················· | ···················· | ···················· | ···················· | ···················· | |
| L | 8943 | hCoV-19/USA/CA-CZB-10907/2020\|EPI_ISL_583136\|2020- | ···················· | ·A·················· | ···················· | ···················· | ···················· | ···················· | |
| L | 8944 | hCoV-19/USA/CA-CZB-10909/2020\|EPI_ISL_583138\|2020- | ···················· | ·A·················· | ···················· | ···················· | ···················· | ···················· | |
| L | 8945 | hCoV-19/USA/CA-CZB-10911/2020\|EPI_ISL_583140\|2020- | ···················· | ·A·················· | ···················· | ···················· | ···················· | ···················· | |
| L | 8946 | hCoV-19/USA/CA-CZB-10912/2020\|EPI_ISL_583141\|2020- | ···················· | ·A·················· | ···················· | ···················· | ···················· | ···················· | |
| L | 8947 | hCoV-19/USA/CA-CZB-10913/2020\|EPI_ISL_583142\|2020- | ···················· | ·A·················· | ···················· | ···················· | ···················· | ···················· | |
| L | 8948 | hCoV-19/USA/CA-CZB-10914/2020\|EPI_ISL_583143\|2020- | ···················· | ·A·················· | ···················· | ···················· | ···················· | ···················· | |
| L | 8949 | hCoV-19/USA/CA-CZB-10915/2020\|EPI_ISL_583144\|2020- | ···················· | ·A·················· | ···················· | ···················· | ···················· | ···················· | |
| L | 8950 | hCoV-19/USA/CA-CZB-10916/2020\|EPI_ISL_583145\|2020- | ···················· | ·A·················· | ···················· | ···················· | ···················· | ···················· | |
| L | 8951 | hCoV-19/USA/CA-CZB-10917/2020\|EPI_ISL_583146\|2020- | ···················· | ·A·················· | ···················· | ···················· | ···················· | ···················· | |
| L | 8952 | hCoV-19/USA/CA-CZB-10918/2020\|EPI_ISL_583147\|2020- | ···················· | ·A·················· | ···················· | ···················· | ···················· | ···················· | |
| L | 8953 | hCoV-19/USA/CA-CZB-10919/2020\|EPI_ISL_583148\|2020- | ···················· | ·A·················· | ···················· | ···················· | ···················· | ···················· | |
| L | 8954 | hCoV-19/USA/CA-CZB-10920/2020\|EPI_ISL_583149\|2020- | ···················· | ·A·················· | ···················· | ···················· | ···················· | ···················· | |
| L | 8955 | hCoV-19/USA/CA-CZB-10921/2020\|EPI_ISL_583150\|2020- | ···················· | ·A·················· | ···················· | ···················· | ···················· | ···················· | |
| L | 8956 | hCoV-19/USA/CA-CZB-10922/2020\|EPI_ISL_583151\|2020- | ···················· | ·A·················· | ···················· | ···················· | ···················· | ···················· | |
| L | 8957 | hCoV-19/USA/CA-CZB-10923/2020\|EPI_ISL_583152\|2020- | ···················· | ·A·················· | ···················· | ···················· | ···················· | ···················· | |
| L | 8958 | hCoV-19/USA/CA-CZB-10924/2020\|EPI_ISL_583153\|2020- | ···················· | ·A·················· | ···················· | ···················· | ···················· | ···················· | |
| L | 8959 | hCoV-19/USA/CA-CZB-10925/2020\|EPI_ISL_583154\|2020- | ···················· | ·A·················· | ···················· | ···················· | ···················· | ···················· | |
| L | 8960 | hCoV-19/USA/CA-CZB-10926/2020\|EPI_ISL_583155\|2020- | ···················· | ···················· | ···················· | ···················· | ···················· | ···················· | |
| L | 8961 | hCoV-19/England/QEUH-9BA901/2020\|EPI_ISL_573610\|20 | ···················· | ···················· | ···················· | ···················· | ···················· | ···················· | |
| L | 8962 | hCoV-19/England/QEUH-9BA93E/2020\|EPI_ISL_573611\|20 | ···················· | ···················· | ···················· | ···················· | ···················· | ···················· | |
| L | 8963 | hCoV-19/England/QEUH-9BA95C/2020\|EPI_ISL_573612\|20 | ···················· | ···················· | ···················· | ···················· | ···················· | ···················· | |
| L | 8964 | hCoV-19/England/QEUH-9BA998/2020\|EPI_ISL_573613\|20 | ···················· | ···················· | ···················· | ···················· | ···················· | ···················· | |
| L | 8965 | hCoV-19/England/QEUH-9BA9A7/2020\|EPI_ISL_573614\|20 | ···················· | ···················· | ···················· | ···················· | ···················· | ···················· | |
| L | 8966 | hCoV-19/England/QEUH-9BA9B6/2020\|EPI_ISL_573615\|20 | ···················· | ·A·················· | ···················· | ···················· | ···················· | ··A················· | |
| L | 8967 | hCoV-19/England/QEUH-9BA9C5/2020\|EPI_ISL_573616\|20 | ···················· | ·A·················· | ···················· | ···················· | ···················· | ···················· | |
| L | 8968 | hCoV-19/England/QEUH-9BA9D4/2020\|EPI_ISL_573617\|20 | ···················· | ···················· | ···················· | ···················· | ···················· | ···················· | |
| L | 8969 | hCoV-19/England/QEUH-9BAA3B/2020\|EPI_ISL_573618\|20 | ···················· | ···················· | ···················· | ···················· | ···················· | ···················· | |
| L | 8970 | hCoV-19/England/QEUH-9BAA4A/2020\|EPI_ISL_573619\|20 | ···················· | ···················· | ···················· | ···················· | ···················· | ···················· | |
| L | 8971 | hCoV-19/England/QEUH-9BAAE0/2020\|EPI_ISL_573620\|20 | ···················· | ···················· | ···················· | ···················· | ···················· | ···················· | |
| L | 8972 | hCoV-19/England/QEUH-9BAB1A/2020\|EPI_ISL_573621\|20 | ···················· | ···················· | ···················· | ···················· | ···················· | ···················· | |
| L | 8973 | hCoV-19/England/QEUH-9BAB29/2020\|EPI_ISL_573622\|20 | ···················· | ···················· | ···················· | ···················· | ···················· | ···················· | |
| L | 8974 | hCoV-19/England/QEUH-9BAB47/2020\|EPI_ISL_573623\|20 | ···················· | ···················· | ···················· | ···················· | ···················· | ···················· | |
| L | 8975 | hCoV-19/England/QEUH-9BAB65/2020\|EPI_ISL_573624\|20 | ···················· | ···················· | ···················· | ···················· | ···················· | ···················· | |
| L | 8976 | hCoV-19/England/QEUH-9BAB74/2020\|EPI_ISL_573625\|20 | ···················· | ···················· | ···················· | ···················· | ···················· | ···················· | |
| L | 8977 | hCoV-19/England/QEUH-9BAB83/2020\|EPI_ISL_573626\|20 | ···················· | ···················· | ···················· | ···················· | ···················· | ···················· | |
| L | 8978 | hCoV-19/England/QEUH-9BAB92/2020\|EPI_ISL_573627\|20 | ···················· | ···················· | ···················· | ···················· | ···················· | ···················· | |
| L | 8979 | hCoV-19/England/QEUH-9BABA1/2020\|EPI_ISL_573628\|20 | ···················· | ·A·················· | ···················· | ···················· | ···················· | ··A················· | |
| L | 8980 | hCoV-19/England/QEUH-9BABFC/2020\|EPI_ISL_573629\|20 | ···················· | ···················· | ···················· | ···················· | ···················· | ···················· | |
| L | 8981 | hCoV-19/Switzerland/BE-ETHZ-100310/2020\|EPI_ISL_57 | ···················· | ···················· | ···················· | ···················· | ···················· | ···················· | |
| L | 8982 | hCoV-19/Switzerland/BE-ETHZ-100340/2020\|EPI_ISL_57 | ···················· | ···················· | ···················· | ···················· | ···················· | ···················· | |
| L | 8983 | hCoV-19/Switzerland/GE-ETHZ-100365/2020\|EPI_ISL_57 | ···················· | ···················· | ···················· | ···················· | ···················· | ···················· | |
| L | 8984 | hCoV-19/Switzerland/GE-ETHZ-100367/2020\|EPI_ISL_57 | ···················· | ···················· | ···················· | ···················· | ···················· | ···················· | |
| L | 8985 | hCoV-19/Switzerland/GE-ETHZ-100368/2020\|EPI_ISL_57 | ···················· | ···················· | ···················· | ···················· | ···················· | ···················· | |
| L | 8986 | hCoV-19/Switzerland/GE-ETHZ-100374/2020\|EPI_ISL_57 | ···················· | ···················A | ···················· | ···················· | ···················· | ···················· | 1 |
| L | 8987 | hCoV-19/Switzerland/GR-ETHZ-100156/2020\|EPI_ISL_57 | ···················· | ···················· | ···················· | ···················· | ···················· | ···················· | |
| L | 8988 | hCoV-19/Switzerland/GR-ETHZ-100157/2020\|EPI_ISL_57 | ···················· | ···················· | ···················· | ···················· | ···················· | ···················· | |
| L | 8989 | hCoV-19/Switzerland/GR-ETHZ-100159/2020\|EPI_ISL_57 | ···················· | ···················· | ···················· | ···················· | ···················· | ···················· | |
| L | 8990 | hCoV-19/Switzerland/GR-ETHZ-100315/2020\|EPI_ISL_57 | ···················· | ···················· | ···················· | ···················· | ···················· | ···················· | |
| L | 8991 | hCoV-19/Switzerland/GR-ETHZ-100319/2020\|EPI_ISL_57 | ···················· | ···················· | ···················· | ···················· | ··············Y····· | ···················· | |
| L | 8992 | hCoV-19/Switzerland/LU-ETHZ-100222/2020\|EPI_ISL_57 | ···················· | ···················· | ···················· | ···················· | ···················· | ···················· | |
| L | 8993 | hCoV-19/Switzerland/LU-ETHZ-100312/2020\|EPI_ISL_57 | ···················· | ···················· | ···················· | ···················· | ···················· | ···················· | |
| L | 8994 | hCoV-19/Switzerland/SZ-ETHZ-100211/2020\|EPI_ISL_57 | ···················· | ···················· | ···················· | ···················· | ···················· | ···················· | |
| L | 8995 | hCoV-19/Switzerland/SZ-ETHZ-100303/2020\|EPI_ISL_57 | ···················· | ···················· | ···················· | ···················· | ···················· | ···················· | |
| L | 8996 | hCoV-19/Switzerland/SZ-ETHZ-100348/2020\|EPI_ISL_57 | ···················· | ···················· | ···················· | ···················· | ···················· | ···················· | |

| | | | | | | | | |
|---|---|---|---|---|---|---|---|---|
| L | 8997 | hCoV-19/Switzerland/SO-ETHZ-100322/2020\|EPI_ISL_57 | .................... | .................... | .................... | .................... | .................... | .................... |
| L | 8998 | hCoV-19/Switzerland/SG-ETHZ-100215/2020\|EPI_ISL_57 | .................... | .................... | .................... | .................... | .................... | .................... |
| L | 8999 | hCoV-19/Switzerland/TG-ETHZ-100239/2020\|EPI_ISL_57 | .................... | .................... | .................... | .................... | .................... | .................... |
| L | 9000 | hCoV-19/Switzerland/TI-ETHZ-100170/2020\|EPI_ISL_57 | .................... | .................... | .................... | .................... | .................... | .................... |
| L | 9001 | hCoV-19/Switzerland/TI-ETHZ-100179/2020\|EPI_ISL_57 | .................... | .................... | .................... | .................... | .................... | .................... |
| L | 9002 | hCoV-19/Switzerland/TI-ETHZ-100182/2020\|EPI_ISL_57 | .................... | .................... | .................... | .................... | .................... | .................... |
| L | 9003 | hCoV-19/England/NORW-F048F/2020\|EPI_ISL_577102\|202 | .................... | .A.................. | .................... | .................... | .................... | ..A................. |
| L | 9004 | hCoV-19/England/NORW-F04AD/2020\|EPI_ISL_577103\|202 | .................... | .................... | .................... | .................... | .................... | .................... |
| L | 9005 | hCoV-19/England/NORW-F0856/2020\|EPI_ISL_577107\|202 | .................... | .................... | .................... | .................... | .................... | .................... |
| L | 9006 | hCoV-19/England/NORW-F0908/2020\|EPI_ISL_577109\|202 | .................... | .................... | .................... | .................... | .................... | .................... |
| L | 9007 | hCoV-19/England/NOTT-113DB7/2020\|EPI_ISL_577110\|20 | .................... | .................... | .................... | .................... | .................... | .................... |
| L | 9008 | hCoV-19/England/NOTT-113DE4/2020\|EPI_ISL_577111\|20 | .................... | .................... | .................... | .................... | .................... | .................... |
| L | 9009 | hCoV-19/England/NOTT-113E0F/2020\|EPI_ISL_577113\|20 | .................... | .................... | .................... | .................... | .................... | .................... |
| L | 9010 | hCoV-19/England/NOTT-113E2D/2020\|EPI_ISL_577114\|20 | .................... | .................... | .................... | .................... | .................... | .................... |
| L | 9011 | hCoV-19/England/NOTT-113E69/2020\|EPI_ISL_577118\|20 | .................... | .................... | .................... | .................... | .................... | .................... |
| L | 9012 | hCoV-19/England/NOTT-113E78/2020\|EPI_ISL_577119\|20 | .................... | .................... | .................... | .................... | .................... | .................... |
| L | 9013 | hCoV-19/England/NOTT-113E96/2020\|EPI_ISL_577120\|20 | .................... | .................... | .................... | .................... | .................... | .................... |
| L | 9014 | hCoV-19/England/NOTT-113EE1/2020\|EPI_ISL_577121\|20 | .................... | .................... | .................... | .................... | .................... | .................... |
| L | 9015 | hCoV-19/England/NOTT-113F1B/2020\|EPI_ISL_577122\|20 | .................... | .................... | .................... | .................... | .................... | .................... |
| L | 9016 | hCoV-19/England/NOTT-113F39/2020\|EPI_ISL_577123\|20 | .................... | .................... | .................... | .................... | .................... | .................... |
| L | 9017 | hCoV-19/England/NOTT-113F84/2020\|EPI_ISL_577124\|20 | .................... | .................... | .................... | .................... | .................... | .................... |
| L | 9018 | hCoV-19/England/NOTT-113F93/2020\|EPI_ISL_577125\|20 | .................... | .................... | .................... | .................... | .................... | .................... |
| L | 9019 | hCoV-19/England/NOTT-113FA2/2020\|EPI_ISL_577126\|20 | .................... | .................... | .................... | .................... | .................... | .................... |
| L | 9020 | hCoV-19/England/NOTT-113FEE/2020\|EPI_ISL_577128\|20 | .................... | .................... | .................... | .................... | .................... | .................... |
| L | 9021 | hCoV-19/England/NOTT-113FFD/2020\|EPI_ISL_577129\|20 | .................... | .................... | .................... | .................... | .................... | .................... |
| L | 9022 | hCoV-19/England/NOTT-114008/2020\|EPI_ISL_577130\|20 | .................... | .A.................. | .................... | .................... | .................... | ..A................. |
| L | 9023 | hCoV-19/England/NOTT-114044/2020\|EPI_ISL_577131\|20 | .................... | .................... | .................... | .................... | .................... | .................... |
| L | 9024 | hCoV-19/England/NOTT-114053/2020\|EPI_ISL_577132\|20 | .................... | .................... | .................... | .................... | .................... | .................... |
| L | 9025 | hCoV-19/England/NOTT-114062/2020\|EPI_ISL_577133\|20 | .................... | .................... | .................... | .................... | .................... | .................... |
| L | 9026 | hCoV-19/England/NOTT-114071/2020\|EPI_ISL_577134\|20 | .................... | .................... | .................... | .................... | .................... | .................... |
| L | 9027 | hCoV-19/England/NOTT-114080/2020\|EPI_ISL_577135\|20 | .................... | .................... | .................... | .................... | .................... | .................... |
| L | 9028 | hCoV-19/England/NOTT-1140CC/2020\|EPI_ISL_577137\|20 | .................... | .................... | .................... | .................... | .................... | .................... |
| L | 9029 | hCoV-19/England/NOTT-114141/2020\|EPI_ISL_577142\|20 | .................... | .................... | .................... | .................... | .................... | .................... |
| L | 9031 | hCoV-19/England/NOTT-11417E/2020\|EPI_ISL_577144\|20 | .................... | .................... | .................... | .................... | .................... | .................... |
| L | 9032 | hCoV-19/England/NOTT-1141AB/2020\|EPI_ISL_577146\|20 | .................... | .................... | .................... | .................... | .................... | .................... |
| L | 9033 | hCoV-19/England/NOTT-1141F6/2020\|EPI_ISL_577149\|20 | .................... | .................... | .................... | .................... | .................... | .................... |
| L | 9034 | hCoV-19/England/NOTT-114202/2020\|EPI_ISL_577150\|20 | .................... | .................... | .................... | .................... | .................... | .................... |
| L | 9035 | hCoV-19/England/NOTT-114220/2020\|EPI_ISL_577151\|20 | .................... | .................... | .................... | .................... | .................... | .................... |
| L | 9036 | hCoV-19/England/NOTT-11425D/2020\|EPI_ISL_577152\|20 | .................... | .................... | .................... | .................... | .................... | .................... |
| L | 9038 | hCoV-19/England/NOTT-1142D5/2020\|EPI_ISL_577156\|20 | .................... | .................... | .................... | .................... | .................... | .................... |
| L | 9039 | hCoV-19/England/NOTT-11431E/2020\|EPI_ISL_577159\|20 | .................... | .................... | .................... | .................... | .................... | .................... |
| L | 9041 | hCoV-19/USA/WI-WSLH-200180/2020\|EPI_ISL_578405\|202 | .................... | .................... | .................... | .................... | .................... | .................... |
| L | 9043 | hCoV-19/USA/WI-WSLH-200182/2020\|EPI_ISL_578407\|202 | .................... | .................... | .................... | .................... | .................... | .................... |
| L | 9044 | hCoV-19/USA/WI-WSLH-200183/2020\|EPI_ISL_578408\|202 | .................... | .................... | .................... | .................... | .................... | .................... |
| L | 9052 | hCoV-19/USA/WI-WSLH-200191/2020\|EPI_ISL_578416\|202 | .................... | .................... | .................... | .................... | .................... | .................... |
| L | 9053 | hCoV-19/USA/WI-WSLH-200193/2020\|EPI_ISL_578417\|202 | .................... | .................... | .................... | .................... | .................... | .................... |
| L | 9054 | hCoV-19/USA/WI-WSLH-200194/2020\|EPI_ISL_578418\|202 | .................... | .................... | .................... | .................... | .................... | .................... |
| L | 9055 | hCoV-19/USA/WI-WSLH-200195/2020\|EPI_ISL_578419\|202 | .................... | .................... | .................... | .................... | .................... | .................... |
| L | 9056 | hCoV-19/USA/WI-WSLH-200196/2020\|EPI_ISL_578420\|202 | .................... | .................... | .................... | .................... | .................... | .................... |
| L | 9071 | hCoV-19/USA/WI-WSLH-200214/2020\|EPI_ISL_578435\|202 | .................... | .................... | .................... | .................... | .................... | .................... |
| L | 9074 | hCoV-19/USA/WI-WSLH-200218/2020\|EPI_ISL_578438\|202 | .................... | .................... | .................... | .................... | .................... | .................... |
| L | 9077 | hCoV-19/USA/WI-WSLH-200221/2020\|EPI_ISL_578441\|202 | .................... | .................... | .................... | .................... | .................... | .................... |
| L | 9081 | hCoV-19/USA/WI-WSLH-200225/2020\|EPI_ISL_578445\|202 | .................... | .................... | .................... | .................... | .................... | .................... |
| L | 9086 | hCoV-19/USA/WI-WSLH-200230/2020\|EPI_ISL_578450\|202 | .................... | .................... | .................... | .................... | .................... | .................... |
| L | 9087 | hCoV-19/USA/WI-WSLH-200231/2020\|EPI_ISL_578451\|202 | .................... | .................... | .................... | .................... | .................... | .................... |
| L | 9089 | hCoV-19/USA/WI-WSLH-200233/2020\|EPI_ISL_578453\|202 | .................... | .................... | .................... | .................... | .................... | .................... |
| L | 9091 | hCoV-19/USA/WI-WSLH-200235/2020\|EPI_ISL_578455\|202 | .................... | .................... | .................... | .................... | .................... | .................... |
| L | 9123 | hCoV-19/England/QEUH-9E4A92/2020\|EPI_ISL_579759\|20 | .................... | .................... | .................... | .................... | .................... | .................... |
| L | 9124 | hCoV-19/England/QEUH-9E4ADE/2020\|EPI_ISL_579760\|20 | .................... | .................... | .................... | .................... | .................... | .................... |
| L | 9125 | hCoV-19/England/QEUH-9E3EF1/2020\|EPI_ISL_579761\|20 | .................... | .................... | .................... | .................... | .................... | .................... |
| L | 9126 | hCoV-19/England/QEUH-9E50AE/2020\|EPI_ISL_579762\|20 | .................... | .................... | .................... | .................... | .................... | .................... |
| L | 9127 | hCoV-19/England/QEUH-9E47D7/2020\|EPI_ISL_579763\|20 | .................... | .................... | .................... | .................... | .................... | .................... |
| L | 9128 | hCoV-19/England/QEUH-9E4CAB/2020\|EPI_ISL_579764\|20 | .................... | .................... | .................... | .................... | .................... | .................... |
| L | 9129 | hCoV-19/England/QEUH-9E4CBA/2020\|EPI_ISL_579765\|20 | .................... | .................... | .................... | .................... | .................... | .................... |
| L | 9130 | hCoV-19/England/QEUH-9E4F75/2020\|EPI_ISL_579766\|20 | .................... | .A.................. | .................... | .................... | .................... | .................... |
| L | 9131 | hCoV-19/England/QEUH-9E49B3/2020\|EPI_ISL_579767\|20 | .................... | .................... | .................... | .................... | ...T................ | .................... |
| L | 9132 | hCoV-19/Northern Ireland/QEUH-9E4E2D/2020\|EPI_ISL_ | .................... | .................... | .................... | .................... | .................... | .................... |
| L | 9133 | hCoV-19/England/QEUH-9E4ED2/2020\|EPI_ISL_579769\|20 | .................... | .................... | .................... | .................... | .................... | .................... |
| L | 9134 | hCoV-19/England/QEUH-9E5633/2020\|EPI_ISL_579770\|20 | .................... | .................... | .................... | .................... | .................... | .................... |
| L | 9135 | hCoV-19/England/QEUH-9E424F/2020\|EPI_ISL_579771\|20 | .................... | .................... | .................... | .................... | .................... | .................... |
| L | 9136 | hCoV-19/England/QEUH-9E4160/2020\|EPI_ISL_579773\|20 | .................... | .................... | .................... | .................... | ......T............. | .................... |
| L | 9137 | hCoV-19/England/QEUH-9E45CE/2020\|EPI_ISL_579774\|20 | .................... | .................... | .................... | .................... | .................... | .................... |
| L | 9138 | hCoV-19/England/QEUH-9E4B9F/2020\|EPI_ISL_579775\|20 | .................... | .................... | .................... | .................... | .................... | .................... |
| L | 9139 | hCoV-19/England/QEUH-9E569D/2020\|EPI_ISL_579776\|20 | .................... | .................... | .................... | .................... | .................... | .................... |
| L | 9140 | hCoV-19/England/QEUH-9E4EC3/2020\|EPI_ISL_579777\|20 | .................... | .................... | .................... | .................... | .................... | .................... |
| L | 9141 | hCoV-19/England/QEUH-9E4C32/2020\|EPI_ISL_579778\|20 | .................... | .................... | .................... | .................... | .................... | .................... |
| L | 9142 | hCoV-19/England/QEUH-9E4F66/2020\|EPI_ISL_579779\|20 | .................... | .................... | .................... | .................... | .................... | .................... |
| L | 9143 | hCoV-19/England/QEUH-9E4397/2020\|EPI_ISL_579780\|20 | .................... | .................... | .................... | .................... | .................... | .................... |
| L | 9144 | hCoV-19/England/QEUH-9E45BF/2020\|EPI_ISL_579781\|20 | .................... | .................... | .................... | .................... | .................... | .................... |
| L | 9145 | hCoV-19/England/QEUH-9E4A83/2020\|EPI_ISL_579782\|20 | .................... | .................... | .................... | .................... | .................... | .................... |

| | | | | | | | | |
|---|---|---|---|---|---|---|---|---|
| L | 9146 | hCoV-19/England/QEUH-9E56BB/2020\|EPI_ISL_579783\|20 | ···················· | ···················· | ···················· | ···················· | ···················· | ···················· |
| L | 9147 | hCoV-19/England/QEUH-9E4C8D/2020\|EPI_ISL_579784\|20 | ···················· | ···················· | ···················· | ···················· | ···················· | ···················· |
| L | 9148 | hCoV-19/England/QEUH-9E4986/2020\|EPI_ISL_579785\|20 | ···················· | ·A·················· | ···················· | ···················· | ···················· | ···A················ |
| L | 9149 | hCoV-19/England/QEUH-9E425E/2020\|EPI_ISL_579786\|20 | ···················· | ·A·················· | ···················· | ···················· | ···················· | ···················· |
| L | 9150 | hCoV-19/England/QEUH-9E4BEA/2020\|EPI_ISL_579787\|20 | ···················· | ···················· | ···················· | ···················· | ······T············· | ···················· |
| L | 9151 | hCoV-19/England/QEUH-9E4977/2020\|EPI_ISL_579788\|20 | ···················· | ···················· | ···················· | ···················· | ···················· | ···················· |
| L | 9152 | hCoV-19/England/QEUH-9E3EC4/2020\|EPI_ISL_579789\|20 | ···················· | ···················· | ···················· | ···················· | ···················· | ···················· |
| L | 9153 | hCoV-19/England/QEUH-9E427C/2020\|EPI_ISL_579790\|20 | ···················· | ···················· | ···················· | ···················· | ···················· | ···················· |
| L | 9154 | hCoV-19/England/QEUH-9E4230/2020\|EPI_ISL_579791\|20 | ···················· | ···················· | ···················· | ···················· | ···················· | ···················· |
| L | 9155 | hCoV-19/England/QEUH-9E491D/2020\|EPI_ISL_579792\|20 | ···················· | ···················· | ···················· | ···················· | ···················· | ···················· |
| L | 9156 | hCoV-19/England/QEUH-9E3F0D/2020\|EPI_ISL_579793\|20 | ···················· | ···················· | ···················· | ···················· | ···················· | ···················· |
| L | 9157 | hCoV-19/England/QEUH-9E4FB1/2020\|EPI_ISL_579795\|20 | ···················· | ···················· | ···················· | ···················· | ···················· | ···················· |
| L | 9158 | hCoV-19/England/QEUH-9E3F58/2020\|EPI_ISL_579796\|20 | ···················· | ···················· | ···················· | ···················· | ···················· | ···················· |
| L | 9159 | hCoV-19/England/QEUH-9E436A/2020\|EPI_ISL_579798\|20 | ···················· | ···················· | ···················· | ···················· | ···················· | ···················· |
| L | 9160 | hCoV-19/England/QEUH-9E4C23/2020\|EPI_ISL_579799\|20 | ···················· | ···················· | ···················· | ···················· | ···················· | ···················· |
| L | 9161 | hCoV-19/England/ALDP-9EDC57/2020\|EPI_ISL_580720\|20 | ···················· | ···················· | ···················· | ···················· | ···················· | ···················· |
| L | 9162 | hCoV-19/England/ALDP-9EDB1E/2020\|EPI_ISL_580722\|20 | ···················· | ·A·················· | ···················· | ···················· | ···················· | ···················· |
| L | 9163 | hCoV-19/England/ALDP-9ED871/2020\|EPI_ISL_580723\|20 | ···················· | ···················· | ···················· | ···················· | ···················· | ···················· |
| L | 9164 | hCoV-19/Wales/MILK-9F03C5/2020\|EPI_ISL_580726\|2020 | ···················· | ···················· | ···················· | ···················· | ···················· | ···················· |
| L | 9165 | hCoV-19/Wales/MILK-9EECED/2020\|EPI_ISL_580728\|2020 | ···················· | ···················· | ···················· | ···················· | ···················· | ···················· |
| L | 9166 | hCoV-19/England/MILK-9F0741/2020\|EPI_ISL_580729\|20 | ···················· | ···················· | ···················· | ···················· | ···················· | ···················· |
| L | 9167 | hCoV-19/England/ALDP-9EDB0F/2020\|EPI_ISL_580730\|20 | ···················· | ·A·················· | ···················· | ···················· | ···················· | ···················· |
| L | 9168 | hCoV-19/England/MILK-9EEA01/2020\|EPI_ISL_580731\|20 | ···················· | ···················· | ···················· | ···················· | ···················· | ···················· |
| L | 9169 | hCoV-19/Wales/MILK-9F05A1/2020\|EPI_ISL_580732\|2020 | ···················· | ···················· | ···················· | ···················· | ···················· | ···················· |
| L | 9170 | hCoV-19/Wales/MILK-9F02E6/2020\|EPI_ISL_580733\|2020 | ···················· | ···················· | ···················· | ···················· | ···················· | ···················· |
| L | 9171 | hCoV-19/England/ALDP-9EDE60/2020\|EPI_ISL_580734\|20 | ···················· | ···················· | ···················· | ···················· | ···················· | ···················· |
| L | 9172 | hCoV-19/England/MILK-9EE97D/2020\|EPI_ISL_580735\|20 | ···················· | ·A·······A·········· | ···················· | ···················· | ···················· | ···················· |
| L | 9173 | hCoV-19/Wales/MILK-9F0671/2020\|EPI_ISL_580736\|2020 | ···················· | ···················· | ···················· | ···················· | ···················· | ···················· |
| L | 9174 | hCoV-19/England/ALDP-9EDC75/2020\|EPI_ISL_580737\|20 | ···················· | ···················· | ···················· | ···················· | ···················· | ···················· |
| L | 9175 | hCoV-19/England/ALDP-9EDA5D/2020\|EPI_ISL_580738\|20 | ···················· | ···················· | ···················· | ···················· | ···················· | ···················· |
| L | 9176 | hCoV-19/England/ALDP-9EDDAF/2020\|EPI_ISL_580740\|20 | ···················· | ···················· | ···················· | ···················· | ···················· | ···················· |
| L | 9177 | hCoV-19/England/MILK-9EE5C4/2020\|EPI_ISL_580741\|20 | ···················· | ···················· | ···················· | ···················· | ···················· | ···················· |
| L | 9178 | hCoV-19/England/MILK-9EE6EF/2020\|EPI_ISL_580742\|20 | ···················· | ···················· | ···················· | ···················· | ···················· | ···················· |
| L | 9179 | hCoV-19/Wales/MILK-9F01BC/2020\|EPI_ISL_580743\|2020 | ···················· | ···················· | ···················· | ···················· | ···················· | ···················· |
| L | 9180 | hCoV-19/England/MILK-9EE8CB/2020\|EPI_ISL_580744\|20 | ···················· | ···················· | ···················· | ···················· | ···················· | ···················· |
| L | 9181 | hCoV-19/Wales/MILK-9F0617/2020\|EPI_ISL_580745\|2020 | ···················· | ···················· | ···················· | ···················· | ···················· | ···················· |
| L | 9182 | hCoV-19/Wales/MILK-9EECCF/2020\|EPI_ISL_580746\|2020 | ···················· | ···················· | ···················· | ···················· | ···················· | ···················· |
| L | 9183 | hCoV-19/England/MILK-9EEE50/2020\|EPI_ISL_580747\|20 | ···················· | ···················· | ···················· | ···················· | ···················· | ···················· |
| L | 9184 | hCoV-19/England/MILK-9F077E/2020\|EPI_ISL_580748\|20 | ···················· | ···················· | ···················· | ···················· | ···················· | ···················· |
| L | 9185 | hCoV-19/England/MILK-9EEC29/2020\|EPI_ISL_580749\|20 | ···················· | ···················· | ···················· | ···················· | ···················· | ···················· |
| L | 9186 | hCoV-19/Wales/MILK-9EE649/2020\|EPI_ISL_580750\|2020 | ···················· | ···················· | ···················· | ···················· | ···················· | ···················· |
| L | 9187 | hCoV-19/Wales/MILK-9EECDE/2020\|EPI_ISL_580752\|2020 | ···················· | ···················· | ···················· | ···················· | ···················· | ···················· |
| L | 9188 | hCoV-19/Wales/MILK-9EEA98/2020\|EPI_ISL_580753\|2020 | ···················· | ···················· | ···················· | ···················· | ···················· | ···················· |
| L | 9189 | hCoV-19/England/ALDP-9ED8DB/2020\|EPI_ISL_580754\|20 | ···················· | ···················· | ···················· | ···················· | ···················· | ···················· |
| L | 9190 | hCoV-19/England/MILK-9EE791/2020\|EPI_ISL_580755\|20 | ···················· | ···················· | ···················· | ···················· | ···················· | ···················· |
| L | 9191 | hCoV-19/England/MILK-9EE63A/2020\|EPI_ISL_580757\|20 | ···················· | ···················· | ···················· | ···················· | ···················· | ···················· |
| L | 9192 | hCoV-19/England/MILK-9F07BA/2020\|EPI_ISL_580760\|20 | ···················· | ···················· | ···················· | ···················· | ···················· | ···················· |
| L | 9193 | hCoV-19/England/MILK-9EEDBD/2020\|EPI_ISL_580762\|20 | ···················· | ···················· | ···················· | ···················· | ···················· | ···················· |
| L | 9194 | hCoV-19/England/ALDP-9EDA7B/2020\|EPI_ISL_580763\|20 | ···················· | ···················· | ···················· | ···················· | ···················· | ···················· |
| L | 9195 | hCoV-19/England/ALDP-9EDD54/2020\|EPI_ISL_580764\|20 | ···················· | ···················· | ···················· | ···················· | ···················· | ···················· |
| L | 9196 | hCoV-19/Wales/MILK-9EE922/2020\|EPI_ISL_580766\|2020 | ···················· | ···················· | ···················· | ···················· | ···················· | ···················· |
| L | 9197 | hCoV-19/England/MILK-9EE3E8/2020\|EPI_ISL_580767\|20 | ···················· | ···················· | ···················· | ···················· | ···················· | ···················· |
| L | 9198 | hCoV-19/Wales/MILK-9F0626/2020\|EPI_ISL_580768\|2020 | ···················· | ···················· | ···················· | ···················· | ···················· | ···················· |
| L | 9199 | hCoV-19/England/ALDP-9ED862/2020\|EPI_ISL_580769\|20 | ···················· | ···················· | ···················· | ···················· | ···················· | ···················· |
| L | 9200 | hCoV-19/England/MILK-9EE7BF/2020\|EPI_ISL_580770\|20 | ···················· | ···················· | ···················· | ···················· | ···················· | ···················· |
| L | 9201 | hCoV-19/England/ALDP-9EDCB1/2020\|EPI_ISL_580771\|20 | ···················· | ···················· | ···················· | ···················· | ···················· | ···················· |
| L | 9202 | hCoV-19/Wales/MILK-9F06F9/2020\|EPI_ISL_580773\|2020 | ···················· | ···················· | ···················· | ···················· | ···················· | ···················· |
| L | 9203 | hCoV-19/England/MILK-9EE500/2020\|EPI_ISL_580775\|20 | ···················· | ···················· | ···················· | ···················· | ···················· | ···················· |
| L | 9204 | hCoV-19/Wales/MILK-9EEA5C/2020\|EPI_ISL_580776\|2020 | ···················· | ···················· | ···················· | ···················· | ···················· | ···················· |
| L | 9205 | hCoV-19/England/MILK-9EE773/2020\|EPI_ISL_580781\|20 | ···················· | ···················· | ···················· | ···················· | ······T············· | ···················· |
| L | 9206 | hCoV-19/England/MILK-9EEB86/2020\|EPI_ISL_580782\|20 | ···················· | ···················· | ···················· | ···················· | ···················· | ···················· |
| L | 9207 | hCoV-19/England/ALDP-9EDC1B/2020\|EPI_ISL_580784\|20 | ···················· | ···················· | ···················· | ···················· | ···················· | ···················· |
| L | 9208 | hCoV-19/England/MILK-9F0802/2020\|EPI_ISL_580786\|20 | ···················· | ···················· | ···················· | ···················· | ···················· | ···················· |
| L | 9209 | hCoV-19/England/ALDP-9ED729/2020\|EPI_ISL_580787\|20 | ···················· | ···················· | ···················· | ···················· | ···················· | ···················· |
| L | 9210 | hCoV-19/England/ALDP-9EDA6C/2020\|EPI_ISL_580788\|20 | ···················· | ···················· | ···················· | ···················· | ···················· | ···················· |
| L | 9211 | hCoV-19/England/ALDP-9EDBC3/2020\|EPI_ISL_580789\|20 | ···················· | ···················· | ···················· | ···················· | ···················· | ···················· |
| L | 9212 | hCoV-19/England/ALDP-9ED96F/2020\|EPI_ISL_580791\|20 | ···················· | ···················· | ···················· | ···················· | ···················· | ···················· |
| L | 9213 | hCoV-19/England/MILK-9EEAC5/2020\|EPI_ISL_580794\|20 | ···················· | ···················· | ···················· | ···················· | ···················· | ···················· |
| L | 9214 | hCoV-19/England/MILK-9F04C2/2020\|EPI_ISL_580795\|20 | ···················· | ···················· | ···················· | ···················· | ···················· | ···················· |
| L | 9215 | hCoV-19/England/MILK-9EE6C1/2020\|EPI_ISL_580796\|20 | ···················· | ·A·················· | ···················· | ···················· | ···················· | ···················· |
| L | 9216 | hCoV-19/Wales/MILK-9EEA3E/2020\|EPI_ISL_580797\|2020 | ···················· | ···················· | ···················· | ···················· | ···················· | ···················· |
| L | 9217 | hCoV-19/USA/MI-QDX-312/2020\|EPI_ISL_571056\|2020-03 | ···················· | ···················· | ···················· | ···················· | ···················· | ···················· |
| L | 9218 | hCoV-19/USA/MI-QDX-313/2020\|EPI_ISL_571057\|2020-03 | ···················· | ···················· | ···················· | ···················· | ···················· | ···················· |
| L | 9219 | hCoV-19/USA/MI-QDX-314/2020\|EPI_ISL_571058\|2020-03 | ···················· | ···················· | ···················· | ···················· | ···················· | ···················· |
| L | 9220 | hCoV-19/USA/MI-QDX-315/2020\|EPI_ISL_571059\|2020-03 | ···················· | ···················· | ···················· | ···················· | ···················· | ···················· |
| L | 9221 | hCoV-19/USA/MI-QDX-316/2020\|EPI_ISL_571060\|2020-03 | ···················· | ···················· | ···················· | ···················· | ···················· | ···················· |
| L | 9222 | hCoV-19/USA/MI-QDX-317/2020\|EPI_ISL_571061\|2020-03 | ···················· | ···················· | ···················· | ···················· | ···················· | ···················· |
| L | 9223 | hCoV-19/USA/MI-QDX-318/2020\|EPI_ISL_571062\|2020-03 | ···················· | ···················· | ···················· | ···················· | ···················· | ···················· |
| L | 9224 | hCoV-19/USA/MI-QDX-319/2020\|EPI_ISL_571063\|2020-03 | ···················· | ···················· | ···················· | ···················· | ···················· | ···················· |
| L | 9225 | hCoV-19/USA/MI-QDX-320/2020\|EPI_ISL_571064\|2020-03 | ···················· | ···················· | ···················· | ···················· | ···················· | ···················· |

| | | | | | | | | |
|---|---|---|---|---|---|---|---|---|
| L | 9226 hCoV-19/USA/MI-QDX-321/2020\|EPI_ISL_571065\|2020-03 | ··················· | ··················· | ···················· | ··················· | ··················· | ··················· | |
| L | 9227 hCoV-19/USA/MI-QDX-322/2020\|EPI_ISL_571066\|2020-03 | ··················· | ··················· | ···················· | ··················· | ··················· | ··················· | |
| L | 9228 hCoV-19/USA/MI-QDX-323/2020\|EPI_ISL_571067\|2020-03 | ··················· | ··················· | ···················· | ··················· | ··················· | ··················· | |
| L | 9229 hCoV-19/USA/MI-QDX-324/2020\|EPI_ISL_571068\|2020-03 | ··················· | ··················· | ···················· | ··················· | ··················· | ··················· | |
| L | 9230 hCoV-19/USA/MI-QDX-325/2020\|EPI_ISL_571069\|2020-03 | ··················· | ··················· | ···················· | ··················· | ··················· | ··················· | |
| L | 9231 hCoV-19/USA/MI-QDX-326/2020\|EPI_ISL_571070\|2020-03 | ··················· | ··················· | ···················· | ··················· | ··················· | ··················· | |
| L | 9232 hCoV-19/USA/MI-QDX-327/2020\|EPI_ISL_571071\|2020-03 | ··················· | ··················· | ···················· | ··················· | ··················· | ··················· | |
| L | 9233 hCoV-19/USA/MI-QDX-328/2020\|EPI_ISL_571072\|2020-03 | ··················· | ··················· | ···················· | ··················· | ··················· | ··················· | |
| L | 9234 hCoV-19/USA/MI-QDX-329/2020\|EPI_ISL_571073\|2020-03 | ··················· | ··················· | ···················· | ··················· | ··················· | ··················· | |
| L | 9235 hCoV-19/USA/MI-QDX-330/2020\|EPI_ISL_571074\|2020-03 | ··················· | ··················· | ···················· | ··················· | ··················· | ··················· | |
| L | 9236 hCoV-19/USA/MI-QDX-331/2020\|EPI_ISL_571075\|2020-03 | ··················· | ··················· | ···················· | ··················· | ··················· | ··················· | |
| L | 9237 hCoV-19/USA/MI-QDX-332/2020\|EPI_ISL_571076\|2020-03 | ··················· | ··················· | ···················· | ··················· | ··················· | ··················· | |
| L | 9238 hCoV-19/USA/MI-QDX-333/2020\|EPI_ISL_571077\|2020-03 | ··················· | ··················· | ···················· | ··················· | ··················· | ··················· | |
| L | 9239 hCoV-19/USA/MI-QDX-334/2020\|EPI_ISL_571078\|2020-03 | ··················· | ··················· | ···················· | ··················· | ··················· | ··················· | |
| L | 9240 hCoV-19/USA/MI-QDX-335/2020\|EPI_ISL_571079\|2020-03 | ··················· | ··················· | ···················· | ··················· | ··················· | ··················· | |
| L | 9241 hCoV-19/USA/MI-QDX-337/2020\|EPI_ISL_571081\|2020-03 | ··················· | ··················· | ···················· | ··················· | ··················· | ··················· | |
| L | 9242 hCoV-19/USA/MI-QDX-338/2020\|EPI_ISL_571082\|2020-03 | ··················· | ··················· | ···················· | ··················· | ··················· | ··················· | |
| L | 9243 hCoV-19/USA/MI-QDX-339/2020\|EPI_ISL_571083\|2020-03 | ··················· | ··················· | ···················· | ··················· | ··················· | ··················· | |
| L | 9244 hCoV-19/USA/MI-QDX-340/2020\|EPI_ISL_571084\|2020-03 | ··················· | ··················· | ···················· | ··················· | ··················· | ··················· | |
| L | 9245 hCoV-19/USA/MI-QDX-341/2020\|EPI_ISL_571085\|2020-03 | ··················· | ··················· | ···················· | ··················· | ··················· | ··················· | |
| L | 9246 hCoV-19/USA/MI-QDX-343/2020\|EPI_ISL_571087\|2020-03 | ··················· | ··················· | ···················· | ··················· | ··················· | ··················· | |
| L | 9247 hCoV-19/USA/MI-QDX-344/2020\|EPI_ISL_571088\|2020-03 | ··················· | ··················· | ···················· | ··················· | ··················· | ··················· | |
| L | 9248 hCoV-19/USA/MI-QDX-345/2020\|EPI_ISL_571089\|2020-03 | ··················· | ··················· | ···················· | ··················· | ··················· | ··················· | |
| L | 9249 hCoV-19/USA/MI-QDX-346/2020\|EPI_ISL_571090\|2020-03 | ··················· | ··················· | ···················· | ··················· | ··················· | ··················· | |
| L | 9250 hCoV-19/USA/MI-QDX-347/2020\|EPI_ISL_571091\|2020-03 | ··················· | ··················· | ···················· | ··················· | ··················· | ··················· | |
| L | 9251 hCoV-19/USA/MI-QDX-348/2020\|EPI_ISL_571092\|2020-03 | ··················· | ··················· | ···················· | ··················· | ··················· | ··················· | |
| L | 9252 hCoV-19/USA/MI-QDX-349/2020\|EPI_ISL_571093\|2020-03 | ··················· | ··················· | ···················· | ··················· | ··················· | ··················· | |
| L | 9253 hCoV-19/USA/MI-QDX-350/2020\|EPI_ISL_571094\|2020-03 | ··················· | ··················· | ···················· | ··················· | ··················· | ··················· | |
| L | 9254 hCoV-19/USA/MI-QDX-351/2020\|EPI_ISL_571095\|2020-03 | ··················· | ··················· | ···················· | ··················· | ··················· | ··················· | |
| L | 9255 hCoV-19/USA/MI-QDX-352/2020\|EPI_ISL_571096\|2020-03 | ··················· | ··················· | ···················· | ··················· | ··················· | ··················· | |
| L | 9256 hCoV-19/USA/MI-QDX-353/2020\|EPI_ISL_571097\|2020-03 | ··················· | ··················· | ···················· | ··················· | ··················· | ··················· | |
| L | 9257 hCoV-19/USA/MI-QDX-354/2020\|EPI_ISL_571098\|2020-03 | ··················· | ··················· | ···················· | ··················· | ··················· | ··················· | |
| L | 9258 hCoV-19/USA/MI-QDX-356/2020\|EPI_ISL_571099\|2020-03 | ··················· | ··················· | ···················· | ··················· | ··················· | ··················· | |
| L | 9259 hCoV-19/USA/VA-DCLS-1441/2020\|EPI_ISL_572308\|2020- | ··················· | ··················· | ···················· | ··················· | ··················· | ··················· | |
| L | 9260 hCoV-19/USA/VA-DCLS-1443/2020\|EPI_ISL_572310\|2020- | ··················· | ··················· | ···················· | ··················· | ··················· | ··················· | |
| L | 9261 hCoV-19/USA/VA-DCLS-1444/2020\|EPI_ISL_572311\|2020- | ··················· | ··················· | ···················· | ··················· | ··················· | ··················· | |
| L | 9262 hCoV-19/USA/VA-DCLS-1445/2020\|EPI_ISL_572312\|2020- | ··················· | ··················· | ···················· | ··················· | ··················· | ··················· | |
| L | 9263 hCoV-19/USA/VA-DCLS-1446/2020\|EPI_ISL_572313\|2020- | ··················· | ··················· | ···················· | ··················· | ··················· | ··················· | |
| L | 9264 hCoV-19/USA/VA-DCLS-1447/2020\|EPI_ISL_572314\|2020- | ··················· | ··················· | ···················· | ··················· | ··················· | ··················· | |
| L | 9265 hCoV-19/USA/VA-DCLS-1448/2020\|EPI_ISL_572315\|2020- | ··················· | ··················· | ···················· | ··················· | ··················· | ··················· | |
| L | 9266 hCoV-19/USA/VA-DCLS-1449/2020\|EPI_ISL_572316\|2020- | ··················· | ··················· | ···················· | ··················· | ··················· | ··················· | |
| L | 9267 hCoV-19/USA/VA-DCLS-1450/2020\|EPI_ISL_572317\|2020- | ··················· | ··················· | ···················· | ··················· | ··················· | ··················· | |
| L | 9268 hCoV-19/USA/VA-DCLS-1451/2020\|EPI_ISL_572318\|2020- | ··················· | ··················· | ···················· | ··················· | ··················· | ··················· | |
| L | 9269 hCoV-19/Italy/CAM-IZSM-112/2020\|EPI_ISL_572320\|202 | ··················· | ··················· | ···················· | ··················· | ··················· | ··················· | |
| L | 9270 hCoV-19/Germany/NW-HHU-D870/2020\|EPI_ISL_572397\|20 | ··················· | ··················· | ···················· | ··················· | ··················· | ··················· | |
| L | 9271 hCoV-19/Russia/Moscow-PMVL-12/2020\|EPI_ISL_572398\| | ··················· | ··················· | ···················· | ··················· | ··················· | ··················· | |
| L | 9272 hCoV-19/England/QEUH-9BAF5A/2020\|EPI_ISL_573640\|20 | ··················· | ·A················· | ···················· | ··················· | ··················· | ··A················ | |
| L | 9273 hCoV-19/England/QEUH-9BAF69/2020\|EPI_ISL_573641\|20 | ··················· | ··················· | ···················· | ··················· | ··················· | ··················· | |
| L | 9274 hCoV-19/England/QEUH-9BAF87/2020\|EPI_ISL_573642\|20 | ··················· | ··················· | ···················· | ··················· | ··················· | ··················· | |
| L | 9275 hCoV-19/England/QEUH-9BAF96/2020\|EPI_ISL_573643\|20 | ··················· | ··················· | ···················· | ··················· | ··················· | ··················· | |
| L | 9276 hCoV-19/England/QEUH-9BAFA5/2020\|EPI_ISL_573644\|20 | ··················· | ··················· | ···················· | ··················· | ···T··············· | ··················· | |
| L | 9277 hCoV-19/England/QEUH-9BB029/2020\|EPI_ISL_573645\|20 | ··················· | ··················· | ···················· | ··················· | ··················· | ··················· | |
| L | 9278 hCoV-19/England/QEUH-9BB038/2020\|EPI_ISL_573646\|20 | ··················· | ·A················· | ···················· | ··················· | ··················· | ··················· | |
| L | 9279 hCoV-19/England/QEUH-9BB056/2020\|EPI_ISL_573647\|20 | ··················· | ··················· | ···················· | ··················· | ··················· | ··················· | |
| L | 9280 hCoV-19/England/QEUH-9BB092/2020\|EPI_ISL_573648\|20 | ··················· | ··················· | ···················· | ··················· | ··················· | ··················· | |
| L | 9281 hCoV-19/England/QEUH-9BB0A1/2020\|EPI_ISL_573649\|20 | ··················· | ··················· | ···················· | ··················· | ··················· | ··················· | |
| L | 9282 hCoV-19/England/QEUH-9BB0CF/2020\|EPI_ISL_573650\|20 | ··················· | ··················· | ···················· | ··················· | ··················· | ··················· | |
| L | 9283 hCoV-19/England/QEUH-9BB0DE/2020\|EPI_ISL_573651\|20 | ··················· | ··················· | ···················· | ··················· | ··················· | ··················· | |
| L | 9284 hCoV-19/Switzerland/VS-ETHZ-100187/2020\|EPI_ISL_57 | ··················· | ··················· | ···················· | ··················· | ··················· | ··················· | |
| L | 9285 hCoV-19/Switzerland/VS-ETHZ-100225/2020\|EPI_ISL_57 | ··················· | ··················· | ···················· | ··················· | ··················· | ··················· | |
| L | 9286 hCoV-19/Switzerland/VS-ETHZ-100326/2020\|EPI_ISL_57 | ··················· | ··················· | ···················· | ··················· | ··················· | ··················· | |
| L | 9287 hCoV-19/Switzerland/VS-ETHZ-100335/2020\|EPI_ISL_57 | ··················· | ··················· | ···················· | ··················· | ··················· | ··················· | |
| L | 9288 hCoV-19/Switzerland/VS-ETHZ-100353/2020\|EPI_ISL_57 | ··················· | ··················· | ···················· | ··················· | ··················· | ··················· | |
| L | 9289 hCoV-19/Switzerland/VS-ETHZ-100360/2020\|EPI_ISL_57 | ··················· | ··················· | ···················· | ··················· | ··················· | ··················· | |
| L | 9290 hCoV-19/Switzerland/VS-ETHZ-100361/2020\|EPI_ISL_57 | ··················· | ··················· | ···················· | ··················· | ··················· | ··················· | |
| L | 9291 hCoV-19/Switzerland/VD-ETHZ-100165/2020\|EPI_ISL_57 | ··················· | ··················· | ···················· | ··················· | ··················· | ··················· | |
| L | 9292 hCoV-19/Switzerland/VD-ETHZ-100167/2020\|EPI_ISL_57 | ··················· | ··················· | ···················· | ··················· | ··················· | ··················· | |
| L | 9293 hCoV-19/Switzerland/VD-ETHZ-100168/2020\|EPI_ISL_57 | ··················· | ··················A | ···················· | ··················· | ··················· | ··················· | 1 |
| L | 9294 hCoV-19/Switzerland/VD-ETHZ-100169/2020\|EPI_ISL_57 | ··················· | ··················· | ···················· | ··················· | ··················· | ··················· | |
| L | 9295 hCoV-19/Switzerland/VD-ETHZ-100171/2020\|EPI_ISL_57 | ··················· | ··················· | ···················· | ··················· | ··················· | ··················· | |
| L | 9296 hCoV-19/Switzerland/VD-ETHZ-100172/2020\|EPI_ISL_57 | ··················· | ··················· | ···················· | ··················· | ··················· | ··················· | |
| L | 9297 hCoV-19/England/NOTT-1140EA/2020\|EPI_ISL_577139\|20 | ··················· | ··················· | ···················· | ··················· | ··················· | ··················· | |
| L | 9298 hCoV-19/England/NOTT-1140F9/2020\|EPI_ISL_577140\|20 | ··················· | ··················· | ···················· | ··················· | ··················· | ··················· | |
| L | 9299 hCoV-19/England/NOTT-114123/2020\|EPI_ISL_577141\|20 | ··················· | ··················· | ···················· | ··················· | ··················· | ··················· | |
| L | 9300 hCoV-19/England/NOTT-11418D/2020\|EPI_ISL_577145\|20 | ··················· | ··················· | ···················· | ··················· | ··················· | ··················· | |
| L | 9303 hCoV-19/USA/WI-WSLH-200319/2020\|EPI_ISL_578500\|202 | ··················· | ··················· | ···················· | ··················· | ··················· | ··················· | |
| L | 9305 hCoV-19/USA/WI-WSLH-200321/2020\|EPI_ISL_578502\|202 | ··················· | ··················· | ···················· | ··················· | ··················· | ··················· | |
| L | 9306 hCoV-19/USA/WI-WSLH-200322/2020\|EPI_ISL_578503\|202 | ··················· | ··················· | ···················· | ··················· | ··················· | ··················· | |
| L | 9307 hCoV-19/USA/WI-WSLH-200323/2020\|EPI_ISL_578504\|202 | ··················· | ··················· | ···················· | ··················· | ··················· | ··················· | |
| L | 9312 hCoV-19/USA/WI-WSLH-200328/2020\|EPI_ISL_578509\|202 | ··················· | ··················· | ···················· | ··················· | ··················· | ··················· | |

| | | | | | | | | |
|---|---|---|---|---|---|---|---|---|
| L | 9313 | hCoV-19/USA/WI-WSLH-200329/2020\|EPI_ISL_578510\|202 | ···················· | ···················· | ····················· | ···················· | ···················· | ···················· |
| L | 9315 | hCoV-19/USA/WI-WSLH-200331/2020\|EPI_ISL_578512\|202 | ···················· | ···················· | ····················· | ···················· | ···················· | ···················· |
| L | 9318 | hCoV-19/USA/WI-WSLH-200340/2020\|EPI_ISL_578515\|202 | ···················· | ···················· | ····················· | ···················· | ···················· | ···················· |
| L | 9323 | hCoV-19/USA/WI-WSLH-200346/2020\|EPI_ISL_578520\|202 | ···················· | ···················· | ····················· | ···················· | ···················· | ···················· |
| L | 9324 | hCoV-19/USA/WI-WSLH-200349/2020\|EPI_ISL_578521\|202 | ···················· | ···················· | ····················· | ···················· | ···················· | ···················· |
| L | 9328 | hCoV-19/England/QEUH-9E4E4B/2020\|EPI_ISL_579800\|20 | ···················· | ···················· | ····················· | ···················· | ···················· | ···················· |
| L | 9329 | hCoV-19/England/QEUH-9E4E1E/2020\|EPI_ISL_579801\|20 | ···················· | ···················· | ····················· | ···················· | ···················· | ···················· |
| L | 9330 | hCoV-19/England/QEUH-9E4ACF/2020\|EPI_ISL_579802\|20 | ···················· | ···················· | ····················· | ···················· | ···················· | ···················· |
| L | 9331 | hCoV-19/England/QEUH-9E3F2B/2020\|EPI_ISL_579803\|20 | ···················· | ···················· | ····················· | ···················· | ···················· | ···················· |
| L | 9332 | hCoV-19/England/QEUH-9E433D/2020\|EPI_ISL_579804\|20 | ···················· | ···················· | ····················· | ···················· | ···················· | ···················· |
| L | 9333 | hCoV-19/England/QEUH-9E4388/2020\|EPI_ISL_579805\|20 | ···················· | ···················· | ····················· | ···················· | ···················· | ···················· |
| L | 9334 | hCoV-19/England/QEUH-9E5554/2020\|EPI_ISL_579806\|20 | ···················· | ···················· | ····················· | ···················· | ···················· | ···················· |
| L | 9335 | hCoV-19/England/QEUH-9E56AC/2020\|EPI_ISL_579807\|20 | ···················· | ···················· | ····················· | ···················· | ···················· | ···················· |
| L | 9336 | hCoV-19/Wales/QEUH-9E5527/2020\|EPI_ISL_579809\|2020 | ···················· | ···················· | ····················· | ···················· | ···················· | ···················· |
| L | 9337 | hCoV-19/England/QEUH-9E49D1/2020\|EPI_ISL_579810\|20 | ···················· | ···················· | ····················· | ···················· | ···················· | ···················· |
| L | 9338 | hCoV-19/England/QEUH-9E426D/2020\|EPI_ISL_579811\|20 | ···················· | ···················· | ····················· | ···················· | ···················· | ···················· |
| L | 9339 | hCoV-19/England/QEUH-9E49FF/2020\|EPI_ISL_579812\|20 | ···················· | ···················· | ····················· | ···················· | ···················· | ···················· |
| L | 9340 | hCoV-19/England/QEUH-9E43A6/2020\|EPI_ISL_579813\|20 | ···················· | ···················· | ····················· | ···················· | ···················· | ···················· |
| L | 9341 | hCoV-19/England/QEUH-9E4E3C/2020\|EPI_ISL_579814\|20 | ···················· | ···················· | ····················· | ···················· | ···················· | ···················· |
| L | 9342 | hCoV-19/England/QEUH-9E479B/2020\|EPI_ISL_579815\|20 | ···················· | ···················· | ····················· | ···················· | ···················· | ···················· |
| L | 9343 | hCoV-19/England/QEUH-9E4959/2020\|EPI_ISL_579816\|20 | ···················· | ···················· | ····················· | ···················· | ···················· | ···················· |
| L | 9344 | hCoV-19/England/QEUH-9E4AA1/2020\|EPI_ISL_579817\|20 | ···················· | ···················· | ····················· | ···················· | ···················· | ···················· |
| L | 9345 | hCoV-19/England/QEUH-9E419D/2020\|EPI_ISL_579818\|20 | ···················· | ···················· | ····················· | ···················· | ···················· | ···················· |
| L | 9346 | hCoV-19/England/QEUH-9E4607/2020\|EPI_ISL_579819\|20 | ···················· | ···················· | ····················· | ···················· | ···················· | ···················· |
| L | 9347 | hCoV-19/England/QEUH-9E567F/2020\|EPI_ISL_579820\|20 | ···················· | ···················· | ····················· | ···················· | ···················· | ···················· |
| L | 9348 | hCoV-19/England/QEUH-9E5642/2020\|EPI_ISL_579821\|20 | ···················· | ···················· | ····················· | ···················· | ···················· | ···················· |
| L | 9349 | hCoV-19/England/QEUH-9E4E0F/2020\|EPI_ISL_579822\|20 | ···················· | ···················· | ····················· | ···················· | ···················· | ···················· |
| L | 9350 | hCoV-19/England/QEUH-9E4A0B/2020\|EPI_ISL_579823\|20 | ···················· | ···················· | ····················· | ···················· | ····T··············· | ···················· |
| L | 9351 | hCoV-19/England/QEUH-9E50CC/2020\|EPI_ISL_579824\|20 | ···················· | ·A·················· | ····················· | ···················· | ···················· | ···A················ |
| L | 9352 | hCoV-19/England/QEUH-9E3F3A/2020\|EPI_ISL_579825\|20 | ···················· | ···················· | ····················· | ···················· | ···················· | ···················· |
| L | 9353 | hCoV-19/England/QEUH-9E45DD/2020\|EPI_ISL_579826\|20 | ···················· | ···················· | ····················· | ···················· | ···················· | ···················· |
| L | 9354 | hCoV-19/England/QEUH-9E4DC6/2020\|EPI_ISL_579827\|20 | ···················· | ···················· | ····················· | ···················· | ···················· | ···················· |
| L | 9355 | hCoV-19/England/QEUH-9E5660/2020\|EPI_ISL_579828\|20 | ···················· | ···················· | ····················· | ···················· | ···················· | ···················· |
| L | 9356 | hCoV-19/England/QEUH-9E4DA8/2020\|EPI_ISL_579829\|20 | ···················· | ···················· | ····················· | ···················· | ···················· | ···················· |
| L | 9357 | hCoV-19/Northern Ireland/QEUH-9E5651/2020\|EPI_ISL_ | ···················· | ···················· | ····················· | ···················· | ···················· | ···················· |
| L | 9358 | hCoV-19/England/QEUH-9E3F67/2020\|EPI_ISL_579831\|20 | ···················· | ···················· | ····················· | ···················· | ···················· | ···················· |
| L | 9359 | hCoV-19/England/QEUH-9E4A1A/2020\|EPI_ISL_579832\|20 | ···················· | ···················· | ····················· | ···················· | ···················· | ···················· |
| L | 9360 | hCoV-19/England/QEUH-9E48C5/2020\|EPI_ISL_579833\|20 | ···················· | ···················· | ····················· | ···················· | ···················· | ···················· |
| L | 9361 | hCoV-19/England/QEUH-9E3EE2/2020\|EPI_ISL_579834\|20 | ···················· | ···················· | ····················· | ···················· | ···················· | ···················· |
| L | 9362 | hCoV-19/England/QEUH-9E4C50/2020\|EPI_ISL_579836\|20 | ···················· | ···················· | ····················· | ···················· | ···················· | ···················· |
| L | 9363 | hCoV-19/England/QEUH-9E494A/2020\|EPI_ISL_579837\|20 | ···················· | ···················· | ····················· | ···················· | ···················· | ···················· |
| L | 9364 | hCoV-19/England/QEUH-9E4BDB/2020\|EPI_ISL_579838\|20 | ···················· | ···················· | ····················· | ···················· | ···················· | ···················· |
| L | 9365 | hCoV-19/England/QEUH-9E47AA/2020\|EPI_ISL_579839\|20 | ···················· | ···················· | ····················· | ···················· | ···················· | ···················· |
| L | 9366 | hCoV-19/England/QEUH-9E4B62/2020\|EPI_ISL_579840\|20 | ···················· | ···················· | ····················· | ···················· | ······T············· | ···················· |
| L | 9367 | hCoV-19/England/QEUH-9E53D2/2020\|EPI_ISL_579841\|20 | ···················· | ···················· | ····················· | ···················· | ···················· | ···················· |
| L | 9368 | hCoV-19/England/QEUH-9E48F2/2020\|EPI_ISL_579842\|20 | ···················· | ···················· | ····················· | ···················· | ···················· | ···················· |
| L | 9369 | hCoV-19/England/QEUH-9E4151/2020\|EPI_ISL_579844\|20 | ···················· | ···················· | ····················· | ···················· | ···················· | ···················· |
| L | 9370 | hCoV-19/England/QEUH-9E5008/2020\|EPI_ISL_579845\|20 | ···················· | ·A·················· | ····················· | ···················· | ···················· | ···················· |
| L | 9371 | hCoV-19/England/QEUH-9E4AED/2020\|EPI_ISL_579846\|20 | ···················· | ···················· | ····················· | ···················· | ···················· | ···················· |
| L | 9372 | hCoV-19/England/QEUH-9E47B9/2020\|EPI_ISL_579847\|20 | ···················· | ···················· | ····················· | ···················· | ···················· | ···················· |
| L | 9373 | hCoV-19/England/QEUH-9E3E97/2020\|EPI_ISL_579848\|20 | ···················· | ···················· | ····················· | ···················· | ···················· | ···················· |
| L | 9374 | hCoV-19/Wales/QEUH-9E490E/2020\|EPI_ISL_579849\|2020 | ···················· | ···················· | ····················· | ···················· | ···················· | ···················· |
| L | 9375 | hCoV-19/England/QEUH-9E43B5/2020\|EPI_ISL_579850\|20 | ···················· | ···················· | ····················· | ···················· | ···················· | ···················· |
| L | 9376 | hCoV-19/England/QEUH-9E535A/2020\|EPI_ISL_579851\|20 | ···················· | ···················· | ····················· | ···················· | ····T··············· | ···················· |
| L | 9377 | hCoV-19/England/QEUH-9E4BF9/2020\|EPI_ISL_579852\|20 | ···················· | ···················· | ····················· | ···················· | ···················· | ···················· |
| L | 9378 | hCoV-19/England/QEUH-9E4F39/2020\|EPI_ISL_579853\|20 | ···················· | ···················· | ····················· | ···················· | ···················· | ···················· |
| L | 9379 | hCoV-19/England/QEUH-9E4AB0/2020\|EPI_ISL_579854\|20 | ···················· | ···················· | ····················· | ···················· | ···················· | ···················· |
| L | 9380 | hCoV-19/England/QEUH-9E4D99/2020\|EPI_ISL_579855\|20 | ···················· | ···················· | ····················· | ···················· | ···················· | ···················· |
| L | 9381 | hCoV-19/England/MILK-9F07F6/2020\|EPI_ISL_580777\|20 | ···················· | ···················· | ····················· | ···················· | ···················· | ···················· |
| L | 9382 | hCoV-19/England/MILK-9EE728/2020\|EPI_ISL_580778\|20 | ···················· | ···················· | ····················· | ···················· | ···················· | ···················· |
| L | 9383 | hCoV-19/England/MILK-9EE51F/2020\|EPI_ISL_580779\|20 | ···················· | ···················· | ····················· | ···················· | ······T············· | ···················· |
| L | 9384 | hCoV-19/England/MILK-9F035C/2020\|EPI_ISL_580800\|20 | ···················· | ···················· | ····················· | ···················· | ···················· | ···················· |
| L | 9385 | hCoV-19/England/MILK-9EE55B/2020\|EPI_ISL_580801\|20 | ···················· | ···················· | ····················· | ···················· | ···················· | ···················· |
| L | 9386 | hCoV-19/England/MILK-9EE834/2020\|EPI_ISL_580802\|20 | ···················· | ···················· | ····················· | ···················· | ···················· | ···················· |
| L | 9387 | hCoV-19/England/MILK-9EECB0/2020\|EPI_ISL_580803\|20 | ···················· | ···················· | ····················· | ···················· | ···················· | ···················· |
| L | 9388 | hCoV-19/England/MILK-9EE9C8/2020\|EPI_ISL_580804\|20 | ···················· | ···················· | ····················· | ···················· | ···················· | ···················· |
| L | 9389 | hCoV-19/England/MILK-9EEF11/2020\|EPI_ISL_580806\|20 | ···················· | ···················· | ····················· | ···················· | ···················· | ···················· |
| L | 9390 | hCoV-19/England/MILK-9EE98C/2020\|EPI_ISL_580807\|20 | ···················· | ···················· | ····················· | ···················· | ···················· | ···················· |
| L | 9391 | hCoV-19/England/MILK-9EE746/2020\|EPI_ISL_580808\|20 | ···················· | ···················· | ····················· | ···················· | ···················· | ···················· |
| L | 9392 | hCoV-19/England/MILK-9EE88F/2020\|EPI_ISL_580809\|20 | ···················· | ···················· | ····················· | ···················· | ···················· | ···················· |
| L | 9393 | hCoV-19/England/MILK-9EE60D/2020\|EPI_ISL_580810\|20 | ···················· | ···················· | ····················· | ···················· | ···················· | ···················· |
| L | 9394 | hCoV-19/Wales/MILK-9F0705/2020\|EPI_ISL_580812\|2020 | ···················· | ···················· | ····················· | ···················· | ···················· | ···················· |
| L | 9395 | hCoV-19/England/MILK-9EE421/2020\|EPI_ISL_580814\|20 | ···················· | ···················· | ····················· | ···················· | ···················· | ···················· |
| L | 9396 | hCoV-19/England/MILK-9EE694/2020\|EPI_ISL_580815\|20 | ···················· | ···················· | ····················· | ···················· | ···················· | ···················· |
| L | 9397 | hCoV-19/England/MILK-9EEE14/2020\|EPI_ISL_580817\|20 | ···················· | ···················· | ····················· | ···················· | ···················· | ···················· |
| L | 9398 | hCoV-19/England/MILK-9F078D/2020\|EPI_ISL_580818\|20 | ···················· | ···················· | ····················· | ···················· | ···················· | ···················· |
| L | 9399 | hCoV-19/Wales/MILK-9F0547/2020\|EPI_ISL_580819\|2020 | ···················· | ···················· | ····················· | ···················· | ···················· | ···················· |
| L | 9400 | hCoV-19/England/MILK-9EE8E9/2020\|EPI_ISL_580820\|20 | ···················· | ···················· | ····················· | ···················· | ···················· | ···················· |
| L | 9401 | hCoV-19/England/ALDP-9ED923/2020\|EPI_ISL_580821\|20 | ···················· | ···················· | ····················· | ···················· | ···················· | ···················· |
| L | 9402 | hCoV-19/England/ALDP-9ED89F/2020\|EPI_ISL_580823\|20 | ···················· | ···················· | ····················· | ···················· | ···················· | ···················· |

| | | | | | | | | |
|---|---|---|---|---|---|---|---|---|
| L | 9403 | hCoV-19/England/MILK-9F0459/2020\|EPI_ISL_580826\|20 | ···················· | ···················· | ···················· | ···················· | ···················· | ···················· |
| L | 9404 | hCoV-19/England/MILK-9F0583/2020\|EPI_ISL_580827\|20 | ···················· | ···················· | ···················· | ···················· | ···················· | ···················· |
| L | 9405 | hCoV-19/England/MILK-9F04FF/2020\|EPI_ISL_580829\|20 | ···················· | ···················· | ···················· | ···················· | ···················· | ···················· |
| L | 9406 | hCoV-19/Wales/MILK-9F06CC/2020\|EPI_ISL_580830\|2020 | ···················· | ···················· | ···················· | ···················· | ···················· | ···················· |
| L | 9407 | hCoV-19/England/MILK-9F02B9/2020\|EPI_ISL_580831\|20 | ···················· | ···················· | ···················· | ···················· | ···················· | ···················· |
| L | 9408 | hCoV-19/England/MILK-9EE588/2020\|EPI_ISL_580832\|20 | ···················· | ···················· | ···················· | ···················· | ···················· | ···················· |
| L | 9409 | hCoV-19/England/ALDP-9EDBF0/2020\|EPI_ISL_580833\|20 | ···················· | ···················· | ···················· | ···················· | ···················· | ···················· |
| L | 9410 | hCoV-19/England/ALDP-9EDB5A/2020\|EPI_ISL_580835\|20 | ···················· | ···················· | ···················· | ···················· | ···················· | ···················· |
| L | 9411 | hCoV-19/England/MILK-9EE412/2020\|EPI_ISL_580836\|20 | ···················· | ···················· | ···················· | ···················· | ···················· | ···················· |
| L | 9412 | hCoV-19/England/MILK-9EE597/2020\|EPI_ISL_580837\|20 | ···················· | ···················· | ···················· | ···················· | ···················· | ···················· |
| L | 9413 | hCoV-19/England/MILK-9F06DB/2020\|EPI_ISL_580839\|20 | ···················· | ·A·················· | ···················· | ···················· | ···················· | ··A················· |
| L | 9414 | hCoV-19/USA/CA-CZB-10951/2020\|EPI_ISL_583035\|2020- | ···················· | ···················· | ···················· | ···················· | ···················· | ···················· |
| L | 9415 | hCoV-19/USA/CA-CZB-10952/2020\|EPI_ISL_583036\|2020- | ···················· | ·A·················· | ···················· | ···················· | ···················· | ···················· |
| L | 9416 | hCoV-19/USA/CA-CZB-10955/2020\|EPI_ISL_583038\|2020- | ···················· | ·A·················· | ···················· | ···················· | ···················· | ···················· |
| L | 9417 | hCoV-19/USA/CA-CZB-10956/2020\|EPI_ISL_583039\|2020- | ···················· | ·A·················· | ···················· | ···················· | ···················· | ···················· |
| L | 9418 | hCoV-19/USA/CA-CZB-10957/2020\|EPI_ISL_583040\|2020- | ···················· | ·A·················· | ···················· | ···················· | ···················· | ···················· |
| L | 9419 | hCoV-19/USA/CA-CZB-10958/2020\|EPI_ISL_583041\|2020- | ···················· | ·A·················· | ···················· | ···················· | ···················· | ···················· |
| L | 9420 | hCoV-19/USA/CA-CZB-10959/2020\|EPI_ISL_583042\|2020- | ···················· | ·A·················· | ···················· | ···················· | ···················· | ···················· |
| L | 9421 | hCoV-19/USA/CA-CZB-10960/2020\|EPI_ISL_583043\|2020- | ···················· | ···················· | ···················· | ···················· | ···················· | ···················· |
| L | 9422 | hCoV-19/USA/CA-CZB-10961/2020\|EPI_ISL_583044\|2020- | ···················· | ·A·················· | ···················· | ···················· | ···················· | ···················· |
| L | 9423 | hCoV-19/USA/CA-CZB-10785/2020\|EPI_ISL_583046\|2020- | ···················· | ···················· | ···················· | ···················· | ···················· | ···················· |
| L | 9424 | hCoV-19/USA/CA-CZB-10786/2020\|EPI_ISL_583047\|2020- | ···················· | ···················· | ···················· | ···················· | ···················· | ···················· |
| L | 9425 | hCoV-19/USA/CA-CZB-10788/2020\|EPI_ISL_583048\|2020- | ···················· | ···················· | ···················· | ···················· | ···················· | ···················· |
| L | 9426 | hCoV-19/USA/CA-CZB-10789/2020\|EPI_ISL_583049\|2020- | ···················· | ···················· | ···················· | ···················· | ···················· | ···················· |
| L | 9427 | hCoV-19/USA/CA-CZB-10790/2020\|EPI_ISL_583050\|2020- | ···················· | ···················· | ···················· | ···················· | ···················· | ···················· |
| L | 9428 | hCoV-19/USA/CA-CZB-10791/2020\|EPI_ISL_583051\|2020- | ···················· | ···················· | ···················· | ···················· | ···················· | ···················· |
| L | 9429 | hCoV-19/USA/CA-CZB-10793/2020\|EPI_ISL_583053\|2020- | ···················· | ···················· | ···················· | ···················· | ···················· | ···················· |
| L | 9430 | hCoV-19/USA/CA-CZB-10794/2020\|EPI_ISL_583054\|2020- | ···················· | ···················· | ···················· | ···················· | ···················· | ···················· |
| L | 9431 | hCoV-19/USA/CA-CZB-10795/2020\|EPI_ISL_583055\|2020- | ···················· | ···················· | ···················· | ···················· | ···················· | ···················· |
| L | 9432 | hCoV-19/USA/CA-CZB-10796/2020\|EPI_ISL_583056\|2020- | ···················· | ···················· | ···················· | ···················· | ···················· | ···················· |
| L | 9433 | hCoV-19/USA/CA-CZB-10797/2020\|EPI_ISL_583057\|2020- | ···················· | ···················· | ···················· | ···················· | ···················· | ···················· |
| L | 9434 | hCoV-19/USA/CA-CZB-10801/2020\|EPI_ISL_583059\|2020- | ···················· | ···················· | ···················· | ···················· | ···················· | ···················· |
| L | 9435 | hCoV-19/USA/CA-CZB-10802/2020\|EPI_ISL_583060\|2020- | ···················· | ···················· | ···················· | ···················· | ···················· | ···················· |
| L | 9436 | hCoV-19/USA/CA-CZB-10804/2020\|EPI_ISL_583061\|2020- | ···················· | ···················· | ···················· | ···················· | ···················· | ···················· |
| L | 9437 | hCoV-19/USA/CA-CZB-10809/2020\|EPI_ISL_583062\|2020- | ···················· | ···················· | ···················· | ···················· | ···················· | ···················· |
| L | 9438 | hCoV-19/USA/CA-CZB-10810/2020\|EPI_ISL_583063\|2020- | ···················· | ···················· | ···················· | ···················· | ···················· | ···················· |
| L | 9439 | hCoV-19/USA/CA-CZB-10811/2020\|EPI_ISL_583064\|2020- | ···················· | ···················· | ···················· | ···················· | ···················· | ···················· |
| L | 9440 | hCoV-19/USA/CA-CZB-10812/2020\|EPI_ISL_583065\|2020- | ···················· | ···················· | ···················· | ···················· | ···················· | ···················· |
| L | 9441 | hCoV-19/USA/CA-CZB-10813/2020\|EPI_ISL_583066\|2020- | ···················· | ···················· | ···················· | ···················· | ···················· | ···················· |
| L | 9442 | hCoV-19/USA/CA-CZB-10818/2020\|EPI_ISL_583068\|2020- | ···················· | ···················· | ···················· | ···················· | ···················· | ···················· |
| L | 9443 | hCoV-19/USA/CA-CZB-10819/2020\|EPI_ISL_583069\|2020- | ···················· | ···················· | ···················· | ···················· | ···················· | ···················· |
| L | 9444 | hCoV-19/USA/CA-CZB-10820/2020\|EPI_ISL_583070\|2020- | ···················· | ·A·················· | ···················· | ···················· | ···················· | ···················· |
| L | 9445 | hCoV-19/USA/CA-CZB-10821/2020\|EPI_ISL_583071\|2020- | ···················· | ·A·················· | ···················· | ···················· | ···················· | ···················· |
| L | 9446 | hCoV-19/USA/CA-CZB-10822/2020\|EPI_ISL_583072\|2020- | ···················· | ·A·················· | ···················· | ···················· | ···················· | ···················· |
| L | 9447 | hCoV-19/USA/CA-CZB-10823/2020\|EPI_ISL_583073\|2020- | ···················· | ·A·················· | ···················· | ···················· | ···················· | ···················· |
| L | 9448 | hCoV-19/USA/CA-CZB-10824/2020\|EPI_ISL_583074\|2020- | ···················· | ·A·················· | ···················· | ···················· | ···················· | ···················· |
| L | 9449 | hCoV-19/USA/CA-CZB-10825/2020\|EPI_ISL_583075\|2020- | ···················· | ·A·················· | ···················· | ···················· | ···················· | ···················· |
| L | 9450 | hCoV-19/USA/CA-CZB-10826/2020\|EPI_ISL_583076\|2020- | ···················· | ···················· | ···················· | ···················· | ···················· | ···················· |
| L | 9451 | hCoV-19/USA/CA-CZB-10828/2020\|EPI_ISL_583078\|2020- | ···················· | ···················· | ···················· | ···················· | ···················· | ···················· |
| L | 9452 | hCoV-19/USA/CA-CZB-10830/2020\|EPI_ISL_583079\|2020- | ···················· | ···················· | ···················· | ···················· | ···················· | ···················· |
| L | 9453 | hCoV-19/USA/CA-CZB-10832/2020\|EPI_ISL_583080\|2020- | ···················· | ···················· | ···················· | ···················· | ···················· | ···················· |
| L | 9454 | hCoV-19/USA/CA-CZB-10834/2020\|EPI_ISL_583081\|2020- | ···················· | ···················· | ···················· | ···················· | ···················· | ···················· |
| L | 9455 | hCoV-19/USA/CA-CZB-10835/2020\|EPI_ISL_583082\|2020- | ···················· | ···················· | ···················· | ···················· | ···················· | ···················· |
| L | 9457 | hCoV-19/USA/CA-CZB-10837/2020\|EPI_ISL_583084\|2020- | ···················· | ···················· | ···················· | ···················· | ···················· | ···················· |
| L | 9458 | hCoV-19/USA/CA-CZB-10839/2020\|EPI_ISL_583085\|2020- | ···················· | ·A·················· | ···················· | ···················· | ···················· | ···················· |
| L | 9459 | hCoV-19/USA/CA-CZB-10841/2020\|EPI_ISL_583086\|2020- | ···················· | ···················· | ···················· | ···················· | ···················· | ···················· |
| L | 9460 | hCoV-19/USA/CA-CZB-10842/2020\|EPI_ISL_583087\|2020- | ···················· | ···················· | ···················· | ···················· | ···················· | ···················· |
| L | 9461 | hCoV-19/USA/CA-CZB-10843/2020\|EPI_ISL_583088\|2020- | ···················· | ···················· | ···················· | ···················· | ···················· | ···················· |
| L | 9462 | hCoV-19/USA/CA-CZB-10844/2020\|EPI_ISL_583089\|2020- | ···················· | ···················· | ···················· | ···················· | ···················· | ···················· |
| L | 9463 | hCoV-19/USA/CA-CZB-10845/2020\|EPI_ISL_583090\|2020- | ···················· | ·A·················· | ···················· | ···················· | ···················· | ···················· |
| L | 9464 | hCoV-19/USA/CA-CZB-10846/2020\|EPI_ISL_583091\|2020- | ···················· | ·A·················· | ···················· | ···················· | ···················· | ···················· |
| L | 9465 | hCoV-19/USA/CA-CZB-10847/2020\|EPI_ISL_583092\|2020- | ···················· | ···················· | ···················· | ···················· | ···················· | ···················· |
| L | 9466 | hCoV-19/USA/CA-CZB-10848/2020\|EPI_ISL_583093\|2020- | ···················· | ·A·················· | ···················· | ···················· | ···················· | ···················· |
| L | 9467 | hCoV-19/USA/CA-CZB-10849/2020\|EPI_ISL_583094\|2020- | ···················· | ···················· | ···················· | ···················· | ···················· | ···················· |
| L | 9468 | hCoV-19/USA/CA-CZB-10850/2020\|EPI_ISL_583095\|2020- | ···················· | ···················· | ···················· | ···················· | ···················· | ···················· |
| L | 9469 | hCoV-19/USA/CA-CZB-10851/2020\|EPI_ISL_583096\|2020- | ···················· | ···················· | ···················· | ···················· | ···················· | ···················· |
| L | 9470 | hCoV-19/USA/CA-CZB-10852/2020\|EPI_ISL_583097\|2020- | ···················· | ···················· | ···················· | ···················· | ···················· | ···················· |
| L | 9471 | hCoV-19/USA/CA-CZB-10854/2020\|EPI_ISL_583098\|2020- | ···················· | ·A·················· | ···················· | ···················· | ···················· | ···················· |
| L | 9472 | hCoV-19/USA/CA-CZB-10855/2020\|EPI_ISL_583099\|2020- | ···················· | ···················· | ···················· | ···················· | ···················· | ···················· |
| L | 9473 | hCoV-19/USA/AL-QDX-394/2020\|EPI_ISL_571016\|2020-03 | ···················· | ···················· | ···················· | ···················· | ···················· | ···················· |
| L | 9474 | hCoV-19/USA/AL-QDX-396/2020\|EPI_ISL_571017\|2020-03 | ···················· | ···················· | ···················· | ···················· | ···················· | ···················· |
| L | 9475 | hCoV-19/USA/AR-QDX-377/2020\|EPI_ISL_571018\|2020-03 | ···················· | ···················· | ···················· | ···················· | ···················· | ···················· |
| L | 9476 | hCoV-19/USA/AZ-QDX-275/2020\|EPI_ISL_571019\|2020-03 | ···················· | ···················· | ···················· | ···················· | ···················· | ···················· |
| L | 9477 | hCoV-19/USA/CA-QDX-285/2020\|EPI_ISL_571020\|2020-03 | ···················· | ···················· | ···················· | ···················· | ···················· | ···················· |
| L | 9478 | hCoV-19/USA/CA-QDX-286/2020\|EPI_ISL_571021\|2020-03 | ···················· | ···················· | ···················· | ···················· | ···················· | ···················· |
| L | 9479 | hCoV-19/USA/CA-QDX-287/2020\|EPI_ISL_571022\|2020-03 | ···················· | ···················· | ···················· | ···················· | ···················· | ···················· |
| L | 9480 | hCoV-19/USA/CA-QDX-289/2020\|EPI_ISL_571023\|2020-03 | ···················· | ···················· | ···················· | ···················· | ···················· | ···················· |
| L | 9481 | hCoV-19/USA/CA-QDX-291/2020\|EPI_ISL_571024\|2020-03 | ···················· | ···················· | ···················· | ···················· | ···················· | ···················· |
| L | 9482 | hCoV-19/USA/CA-QDX-292/2020\|EPI_ISL_571025\|2020-03 | ···················· | ···················· | ···················· | ···················· | ···················· | ···················· |
| L | 9483 | hCoV-19/USA/CA-QDX-293/2020\|EPI_ISL_571026\|2020-03 | ···················· | ···················· | ···················· | ···················· | ···················· | ···················· |

```
L 9484 hCoV-19/USA/FL-QDX-272/2020|EPI_ISL_571027|2020-03 ···················· ···················· ····················· ···················· ···················· ····················
L 9485 hCoV-19/USA/FL-QDX-273/2020|EPI_ISL_571028|2020-03 ···················· ···················· ····················· ···················· ···················· ····················
L 9486 hCoV-19/USA/FL-QDX-278/2020|EPI_ISL_571029|2020-03 ···················· ···················· ····················· ···················· ···················· ····················
L 9487 hCoV-19/USA/IL-QDX-279/2020|EPI_ISL_571030|2020-03 ···················· ···················· ····················· ···················· ···················· ····················
L 9488 hCoV-19/USA/IL-QDX-280/2020|EPI_ISL_571031|2020-03 ···················· ···················· ····················· ···················· ···················· ····················
L 9489 hCoV-19/USA/IL-QDX-282/2020|EPI_ISL_571033|2020-03 ···················· ···················· ····················· ···················· ···················· ····················
L 9490 hCoV-19/USA/IL-QDX-283/2020|EPI_ISL_571034|2020-03 ···················· ···················· ····················· ···················· ···················· ····················
L 9491 hCoV-19/USA/IL-QDX-284/2020|EPI_ISL_571035|2020-03 ···················· ···················· ····················· ···················· ···················· ····················
L 9492 hCoV-19/USA/MI-QDX-274/2020|EPI_ISL_571036|2020-03 ···················· ·············A······ ····················· ···················· ···················· ····················
L 9493 hCoV-19/USA/MI-QDX-277/2020|EPI_ISL_571037|2020-03 ···················· ···················· ····················· ···················· ···················· ····················
L 9494 hCoV-19/USA/MI-QDX-294/2020|EPI_ISL_571038|2020-03 ···················· ···················· ····················· ···················· ···················· ····················
L 9495 hCoV-19/USA/MI-QDX-295/2020|EPI_ISL_571039|2020-03 ···················· ···················· ····················· ···················· ···················· ····················
L 9496 hCoV-19/USA/MI-QDX-296/2020|EPI_ISL_571040|2020-03 ···················· ···················· ····················· ···················· ···················· ····················
L 9497 hCoV-19/USA/MI-QDX-297/2020|EPI_ISL_571041|2020-03 ···················· ···················· ····················· ···················· ···················· ····················
L 9498 hCoV-19/USA/MI-QDX-298/2020|EPI_ISL_571042|2020-03 ···················· ···················· ····················· ···················· ···················· ····················
L 9499 hCoV-19/USA/MI-QDX-299/2020|EPI_ISL_571043|2020-03 ···················· ···················· ····················· ···················· ···················· ····················
L 9500 hCoV-19/USA/MI-QDX-300/2020|EPI_ISL_571044|2020-03 ···················· ···················· ····················· ···················· ···················· ····················
L 9501 hCoV-19/USA/MI-QDX-301/2020|EPI_ISL_571045|2020-03 ···················· ···················· ····················· ···················· ···················· ····················
L 9502 hCoV-19/USA/MI-QDX-302/2020|EPI_ISL_571046|2020-03 ···················· ···················· ····················· ···················· ···················· ····················
L 9503 hCoV-19/USA/MI-QDX-303/2020|EPI_ISL_571047|2020-03 ···················· ···················· ····················· ···················· ···················· ····················
L 9504 hCoV-19/USA/MI-QDX-304/2020|EPI_ISL_571048|2020-03 ···················· ···················· ····················· ···················· ···················· ····················
L 9505 hCoV-19/USA/MI-QDX-305/2020|EPI_ISL_571049|2020-03 ···················· ···················· ····················· ···················· ···················· ····················
L 9506 hCoV-19/USA/MI-QDX-306/2020|EPI_ISL_571050|2020-03 ···················· ···················· ····················· ···················· ···················· ····················
L 9507 hCoV-19/USA/MI-QDX-307/2020|EPI_ISL_571051|2020-03 ···················· ···················· ····················· ···················· ···················· ····················
L 9508 hCoV-19/USA/MI-QDX-308/2020|EPI_ISL_571052|2020-03 ···················· ···················· ····················· ···················· ···················· ····················
L 9509 hCoV-19/USA/MI-QDX-309/2020|EPI_ISL_571053|2020-03 ···················· ···················· ····················· ···················· ···················· ····················
L 9510 hCoV-19/USA/MI-QDX-310/2020|EPI_ISL_571054|2020-03 ···················· ···················· ····················· ···················· ···················· ····················
L 9511 hCoV-19/USA/MI-QDX-311/2020|EPI_ISL_571055|2020-03 ···················· ···················· ····················· ···················· ···················· ····················
L 9512 hCoV-19/USA/VA-DCLS-1286/2020|EPI_ISL_572300|2020- ···················· ···················· ····················· ···················· ···················· ····················
L 9513 hCoV-19/USA/VA-DCLS-1287/2020|EPI_ISL_572301|2020- ···················· ···················· ····················· ···················· ···················· ····················
L 9514 hCoV-19/USA/VA-DCLS-1288/2020|EPI_ISL_572302|2020- ···················· ···················· ····················· ···················· ···················· ····················
L 9515 hCoV-19/USA/VA-DCLS-1289/2020|EPI_ISL_572303|2020- ···················· ···················· ····················· ···················· ···················· ····················
L 9516 hCoV-19/USA/VA-DCLS-1437/2020|EPI_ISL_572304|2020- ···················· ···················· ····················· ···················· ···················· ····················
L 9517 hCoV-19/USA/VA-DCLS-1438/2020|EPI_ISL_572305|2020- ···················· ···················· ····················· ···················· ···················· ····················
L 9518 hCoV-19/USA/VA-DCLS-1439/2020|EPI_ISL_572306|2020- ···················· ···················· ····················· ···················· ···················· ····················
L 9519 hCoV-19/USA/VA-DCLS-1440/2020|EPI_ISL_572307|2020- ···················· ···················· ····················· ···················· ···················· ····················
L 9520 hCoV-19/USA/VA-DCLS-1442/2020|EPI_ISL_572309|2020- ···················· ···················· ····················· ···················· ···················· ····················
L 9521 hCoV-19/Italy/CAM-IZSM-129/2020|EPI_ISL_572321|202 ···················· ···················· ····················· ···················· ···················· ····················
L 9522 hCoV-19/Italy/CAM-IZSM-166/2020|EPI_ISL_572322|202 ··················T· ···················· ····················· ···················· ···················· ···················· 1
L 9523 hCoV-19/Italy/CAM-IZSM-172/2020|EPI_ISL_572323|202 ···················· ···················· ····················· ···················· ···················· ····················
L 9524 hCoV-19/Italy/CAM-IZSM-32830/2020|EPI_ISL_572324|2 ···················· ···················· ····················· ···················· ···················· ····················
L 9525 hCoV-19/Slovakia/UKBA-101/2020|EPI_ISL_572329|2020 ···················· ···················· ····················· ···················· ···················· ····················
L 9526 hCoV-19/Germany/NW-HHU-D808/2020|EPI_ISL_572330|20 ···················· ···················· ····················· ···················· ···················· ····················
L 9527 hCoV-19/Germany/NW-HHU-D809/2020|EPI_ISL_572331|20 ···················· ···················· ····················· ···················· ···················· ····················
L 9528 hCoV-19/Brazil/PE-IAM109/2020|EPI_ISL_572334|2020- ···················· ···················· ····················· ···················· ···················· ····················
L 9529 hCoV-19/Brazil/PE-IAM383/2020|EPI_ISL_572335|2020- ···················· ···················· ····················· ···················· ···················· ····················
L 9530 hCoV-19/Brazil/PE-IAM711/2020|EPI_ISL_572351|2020- ···················· ···················· ····················· ···················· ···················· ····················
L 9531 hCoV-19/Brazil/PE-IAM719/2020|EPI_ISL_572353|2020- ···················· ···················· ····················· ···················· ···················· ····················
L 9532 hCoV-19/Brazil/PE-IAM961/2020|EPI_ISL_572366|2020- ···················· ···················· ····················· ···················· ···················· ····················
L 9533 hCoV-19/Brazil/PE-IAM991/2020|EPI_ISL_572371|2020- ···················· ···················· ····················· ···················· ···················· ····················
L 9534 hCoV-19/Brazil/PE-IAM1309/2020|EPI_ISL_572386|2020 ···················· ···················· ····················· ···················· ···················· ····················
L 9535 hCoV-19/England/QEUH-9BAC08/2020|EPI_ISL_573630|20 ···················· ···················· ····················· ···················· ···················· ····················
L 9536 hCoV-19/England/QEUH-9BAC53/2020|EPI_ISL_573631|20 ···················· ···················· ····················· ···················· ···················· ····················
L 9537 hCoV-19/England/QEUH-9BACAE/2020|EPI_ISL_573632|20 ···················· ···················· ····················· ···················· ···················· ····················
L 9538 hCoV-19/England/QEUH-9BACCC/2020|EPI_ISL_573633|20 ···················· ·A·················· ····················· ···················· ···················· ···A················
L 9539 hCoV-19/England/QEUH-9BACEA/2020|EPI_ISL_573634|20 ···················· ···················· ····················· ···················· ···················· ····················
L 9540 hCoV-19/England/QEUH-9BACF9/2020|EPI_ISL_573635|20 ···················· ···················· ····················· ···················· ···················· ····················
L 9541 hCoV-19/England/QEUH-9BAD05/2020|EPI_ISL_573636|20 ···················· ·A·················· ····················· ···················· ···················· ····················
L 9542 hCoV-19/England/QEUH-9BAD14/2020|EPI_ISL_573637|20 ···················· ···················· ····················· ···················· ···················· ····················
L 9543 hCoV-19/England/QEUH-9BAD32/2020|EPI_ISL_573638|20 ···················· ···················· ····················· ···················· ···················· ····················
L 9544 hCoV-19/England/QEUH-9BAF4B/2020|EPI_ISL_573639|20 ···················· ···················· ····················· ···················· ···················· ····················
L 9545 hCoV-19/England/QEUH-9D1694/2020|EPI_ISL_573652|20 ···················· ···················· ····················· ···················· ···················· ····················
L 9546 hCoV-19/England/QEUH-9D16B2/2020|EPI_ISL_573653|20 ···················· ···················· ····················· ···················· ···················· ····················
L 9547 hCoV-19/England/QEUH-9D16C1/2020|EPI_ISL_573654|20 ···················· ···················· ····················· ···················· ···················· ····················
L 9548 hCoV-19/England/QEUH-9D16D0/2020|EPI_ISL_573655|20 ···················· ·A·················· ····················· ···················· ···················· ····················
L 9549 hCoV-19/England/QEUH-9D1737/2020|EPI_ISL_573656|20 ···················· ···················· ····················· ···················· ····T··············· ····················
L 9550 hCoV-19/England/QEUH-9D1746/2020|EPI_ISL_573657|20 ···················· ···················· ····················· ···················· ···················· ····················
L 9551 hCoV-19/England/QEUH-9D1791/2020|EPI_ISL_573658|20 ···················· ···················· ····················· ···················· ···················· ····················
L 9552 hCoV-19/England/QEUH-9D17A0/2020|EPI_ISL_573659|20 ···················· ···················· ····················· ···················· ···················· ····················
L 9553 hCoV-19/England/QEUH-9D17EC/2020|EPI_ISL_573660|20 ···················· ···················· ····················· ···················· ···················· ····················
L 9554 hCoV-19/England/QEUH-9D1807/2020|EPI_ISL_573661|20 ···················· ···················· ····················· ···················· ···················· ····················
L 9555 hCoV-19/England/QEUH-9D1843/2020|EPI_ISL_573662|20 ···················· ···················· ····················· ···················· ···················· ····················
L 9556 hCoV-19/England/QEUH-9D1852/2020|EPI_ISL_573663|20 ···················· ···················· ····················· ···················· ···················· ····················
L 9557 hCoV-19/England/QEUH-9D1861/2020|EPI_ISL_573664|20 ···················· ···················· ····················· ···················· ···················· ····················
L 9558 hCoV-19/England/QEUH-9D1870/2020|EPI_ISL_573665|20 ···················· ···················· ····················· ···················· ····T··············· ····················
L 9559 hCoV-19/England/QEUH-9D188F/2020|EPI_ISL_573666|20 ···················· ···················· ····················· ···················· ···················· ····················
L 9560 hCoV-19/England/QEUH-9D18CB/2020|EPI_ISL_573667|20 ···················· ···················· ····················· ···················· ···················· ····················
L 9561 hCoV-19/England/QEUH-9D18DA/2020|EPI_ISL_573668|20 ···················· ···················· ····················· ···················· ···················· ····················
L 9562 hCoV-19/England/QEUH-9D18E9/2020|EPI_ISL_573669|20 ···················· ···················· ····················· ···················· ····T··············· ····················
L 9563 hCoV-19/England/QEUH-9D1904/2020|EPI_ISL_573670|20 ···················· ···················· ····················· ···················· ···················· ····················
```

| | | | | | | | | |
|---|---|---|---|---|---|---|---|---|
| L | 9564 hCoV-19/England/QEUH-9D19AA/2020\|EPI_ISL_573671\|20 | .................... | .................... | .................... | .................... | ......N............. | .................... | |
| L | 9565 hCoV-19/England/QEUH-9D19B9/2020\|EPI_ISL_573672\|20 | .................... | .................... | .................... | .................... | .................... | .................... | |
| L | 9566 hCoV-19/England/QEUH-9D19C8/2020\|EPI_ISL_573673\|20 | .................... | .................... | .................... | .................... | .................... | .................... | |
| L | 9567 hCoV-19/England/QEUH-9D19E6/2020\|EPI_ISL_573674\|20 | .................... | .................... | .................... | .................... | .................... | .................... | |
| L | 9568 hCoV-19/England/QEUH-9D19F5/2020\|EPI_ISL_573675\|20 | .................... | .................... | .................... | .................... | .................... | .................... | |
| L | 9569 hCoV-19/England/QEUH-9D1A01/2020\|EPI_ISL_573676\|20 | .................... | .................... | .................... | .................... | .................... | .................... | |
| L | 9570 hCoV-19/England/QEUH-9D1A10/2020\|EPI_ISL_573677\|20 | .................... | .................... | .................... | .................... | .................... | .................... | |
| L | 9571 hCoV-19/England/QEUH-9D1A2F/2020\|EPI_ISL_573678\|20 | .................... | .A.................. | .................... | .................... | .................... | .................... | |
| L | 9572 hCoV-19/England/QEUH-9D1A3E/2020\|EPI_ISL_573679\|20 | .................... | .................... | .................... | .................... | .................... | .................... | |
| L | 9573 hCoV-19/England/QEUH-9D1B0E/2020\|EPI_ISL_573680\|20 | .................... | .................... | .................... | .................... | .................... | .................... | |
| L | 9574 hCoV-19/England/QEUH-9D1B1D/2020\|EPI_ISL_573681\|20 | .................... | .................... | .................... | .................... | .................... | .................... | |
| L | 9575 hCoV-19/England/QEUH-9D1B77/2020\|EPI_ISL_573682\|20 | .................... | .................... | .................... | .................... | .................... | .................... | |
| L | 9576 hCoV-19/England/QEUH-9D1C83/2020\|EPI_ISL_573684\|20 | .................... | .................... | .................... | .................... | .................... | .................... | |
| L | 9577 hCoV-19/England/QEUH-9D1C92/2020\|EPI_ISL_573685\|20 | .................... | .................... | .................... | .................... | .................... | .................... | |
| L | 9578 hCoV-19/England/SHEF-C09B6/2020\|EPI_ISL_573686\|202 | .................... | .................... | .................... | .................... | .................... | .................... | |
| L | 9579 hCoV-19/England/SHEF-C0AFF/2020\|EPI_ISL_573687\|202 | .................... | .................... | .................... | .................... | .................... | .................... | |
| L | 9580 hCoV-19/England/SHEF-C12F6/2020\|EPI_ISL_573688\|202 | .................... | .................... | .................... | .................... | .................... | .................... | |
| L | 9581 hCoV-19/England/SHEF-C276F/2020\|EPI_ISL_573689\|202 | .................... | .................... | .................... | .................... | .................... | .................... | |
| L | 9582 hCoV-19/England/SHEF-C2C15/2020\|EPI_ISL_573690\|202 | .................... | .................... | .................... | .................... | .................... | .................... | |
| L | 9583 hCoV-19/England/SHEF-C2D4F/2020\|EPI_ISL_573691\|202 | .................... | .................... | .................... | .................... | .................... | .................... | |
| L | 9584 hCoV-19/England/SHEF-C2F94/2020\|EPI_ISL_573692\|202 | .................... | .................... | .................... | .................... | .................... | .................... | |
| L | 9585 hCoV-19/England/SHEF-C30EB/2020\|EPI_ISL_573693\|202 | .................... | .................... | .................... | .................... | .................... | .................... | |
| L | 9586 hCoV-19/England/SHEF-C30FA/2020\|EPI_ISL_573694\|202 | .................... | .................... | .................... | .................... | .................... | .................... | |
| L | 9587 hCoV-19/England/SHEF-C3731/2020\|EPI_ISL_573695\|202 | .................... | .................... | .................... | .................... | .................... | .................... | |
| L | 9588 hCoV-19/England/SHEF-C37AA/2020\|EPI_ISL_573696\|202 | .................... | .................... | .................... | .................... | .................... | .................... | |
| L | 9589 hCoV-19/England/SHEF-C3D02/2020\|EPI_ISL_573697\|202 | .................... | .................... | .................... | .................... | .................... | .................... | |
| L | 9590 hCoV-19/England/SHEF-C44B1/2020\|EPI_ISL_573698\|202 | .................... | .................... | .................... | .................... | .................... | .................... | |
| L | 9591 hCoV-19/England/SHEF-C4B34/2020\|EPI_ISL_573699\|202 | .................... | .A.................. | .................... | .................... | .................... | .................... | |
| L | 9592 hCoV-19/Switzerland/TI-ETHZ-100185/2020\|EPI_ISL_57 | .................... | .................... | .................... | .................... | .................... | ......M............. | |
| L | 9593 hCoV-19/Switzerland/TI-ETHZ-100221/2020\|EPI_ISL_57 | .................... | .................... | .................... | .................... | .................... | .................... | |
| L | 9594 hCoV-19/Switzerland/TI-ETHZ-100223/2020\|EPI_ISL_57 | .................... | .................... | .................... | .................... | .................... | .................... | |
| L | 9595 hCoV-19/Switzerland/TI-ETHZ-100228/2020\|EPI_ISL_57 | .................... | .................... | .................... | .................... | .................... | .................... | |
| L | 9596 hCoV-19/Switzerland/TI-ETHZ-100337/2020\|EPI_ISL_57 | .................... | .................... | .................... | .................... | .................... | .................... | |
| L | 9597 hCoV-19/Switzerland/UR-ETHZ-100227/2020\|EPI_ISL_57 | .................... | .................... | .................... | .................... | .................... | .................... | |
| L | 9598 hCoV-19/Switzerland/UR-ETHZ-100233/2020\|EPI_ISL_57 | .................... | .................... | .................... | .................... | .................... | .................... | |
| L | 9599 hCoV-19/Switzerland/UR-ETHZ-100321/2020\|EPI_ISL_57 | .................... | .................... | .................... | .................... | .................... | .................... | |
| L | 9600 hCoV-19/Switzerland/VS-ETHZ-100173/2020\|EPI_ISL_57 | .................... | .................... | .................... | .................... | .................... | .................... | |
| L | 9601 hCoV-19/Switzerland/VD-ETHZ-100174/2020\|EPI_ISL_57 | .................... | .A.................. | .................... | .................... | .................... | .................... | |
| L | 9602 hCoV-19/Switzerland/VD-ETHZ-100176/2020\|EPI_ISL_57 | .................... | .................... | .................... | .................... | .................... | .................... | |
| L | 9603 hCoV-19/Switzerland/VD-ETHZ-100178/2020\|EPI_ISL_57 | .................... | .................... | .................... | .................... | .................... | .................... | |
| L | 9604 hCoV-19/Switzerland/VD-ETHZ-100180/2020\|EPI_ISL_57 | .................... | .................... | .................... | .................... | .................... | .................... | |
| L | 9605 hCoV-19/Switzerland/VD-ETHZ-100181/2020\|EPI_ISL_57 | .................... | .A.................. | .................... | .................... | .................... | .................... | |
| L | 9606 hCoV-19/Switzerland/VD-ETHZ-100184/2020\|EPI_ISL_57 | .................... | .................... | .................... | .................... | .................... | .................... | |
| L | 9607 hCoV-19/Switzerland/VD-ETHZ-100186/2020\|EPI_ISL_57 | .................... | .................... | .................... | .................... | .................... | .................... | |
| L | 9608 hCoV-19/Switzerland/VD-ETHZ-100192/2020\|EPI_ISL_57 | .................... | .................... | .................... | .................... | .................... | .................... | |
| L | 9609 hCoV-19/Switzerland/VD-ETHZ-100196/2020\|EPI_ISL_57 | .................... | .................... | .................... | .................... | .................... | .................... | |
| L | 9610 hCoV-19/Switzerland/VD-ETHZ-100198/2020\|EPI_ISL_57 | ........Y........... | .R.................. | .................... | .................... | .................... | .................... | |
| L | 9611 hCoV-19/Switzerland/VD-ETHZ-100200/2020\|EPI_ISL_57 | .................... | .................... | .................... | .................... | .................... | .................... | |
| L | 9612 hCoV-19/Switzerland/VD-ETHZ-100220/2020\|EPI_ISL_57 | .................... | .R.................. | .................... | .................... | .................... | .................... | |
| L | 9613 hCoV-19/Switzerland/VD-ETHZ-100229/2020\|EPI_ISL_57 | .................... | ...................A | .................... | .................... | .................... | .................... | 1 |
| L | 9614 hCoV-19/Switzerland/VD-ETHZ-100230/2020\|EPI_ISL_57 | .................... | .................... | .................... | .................... | .................... | .................... | |
| L | 9615 hCoV-19/Switzerland/VD-ETHZ-100231/2020\|EPI_ISL_57 | .................... | .A.................. | .................... | .................... | .................... | .................... | |
| L | 9616 hCoV-19/Switzerland/VD-ETHZ-100232/2020\|EPI_ISL_57 | .................... | ...................A | .................... | .................... | .................... | .................... | 1 |
| L | 9617 hCoV-19/USA/CA-CZB-6747/2020\|EPI_ISL_583000\|2020-0 | .................... | .A.................. | .................... | .................... | .................... | .................... | |
| L | 9618 hCoV-19/USA/CA-CZB-215/2020\|EPI_ISL_583001\|2020-08 | .................... | .................... | .................... | .................... | .................... | .................... | |
| L | 9619 hCoV-19/USA/CA-CZB-216/2020\|EPI_ISL_583002\|2020-08 | .................... | .................... | .................... | .................... | .................... | .................... | |
| L | 9620 hCoV-19/USA/CA-CZB-218/2020\|EPI_ISL_583003\|2020-08 | .................... | .A.................. | .................... | .................... | .................... | .................... | |
| L | 9621 hCoV-19/USA/CA-CZB-219/2020\|EPI_ISL_583004\|2020-08 | .................... | .A.................. | .................... | .................... | .................... | .................... | |
| L | 9622 hCoV-19/USA/CA-CZB-230/2020\|EPI_ISL_583006\|2020-08 | .................... | .................... | .................... | .................... | .................... | .................... | |
| L | 9623 hCoV-19/USA/CA-CZB-231/2020\|EPI_ISL_583007\|2020-08 | .................... | .................... | .................... | .................... | .................... | .................... | |
| L | 9624 hCoV-19/USA/CA-CZB-7128/2020\|EPI_ISL_583008\|2020-0 | .................... | .................... | .................... | .................... | .................... | .................... | |
| L | 9625 hCoV-19/USA/CA-CZB-7133/2020\|EPI_ISL_583011\|2020-0 | .................... | .A.................. | .................... | .................... | .................... | .................... | |
| L | 9626 hCoV-19/USA/CA-CZB-7134/2020\|EPI_ISL_583012\|2020-0 | .................... | .A.................. | .................... | .................... | .................... | .................... | |
| L | 9627 hCoV-19/USA/CA-CZB-7135/2020\|EPI_ISL_583013\|2020-0 | .................... | .A.................. | .................... | .................... | .................... | .................... | |
| L | 9628 hCoV-19/USA/CA-CZB-7139/2020\|EPI_ISL_583014\|2020-0 | .................... | .A.................. | .................... | .................... | .................... | .................... | |
| L | 9629 hCoV-19/USA/CA-CZB-7140/2020\|EPI_ISL_583015\|2020-0 | .................... | .A.................. | .................... | .................... | .................... | .................... | |
| L | 9630 hCoV-19/USA/CA-CZB-10285/2020\|EPI_ISL_583016\|2020- | .................... | .................... | .................... | .................... | .................... | .................... | |
| L | 9631 hCoV-19/USA/CA-CZB-10287/2020\|EPI_ISL_583017\|2020- | .................... | .................... | .................... | .................... | .................... | .................... | |
| L | 9632 hCoV-19/USA/CA-CZB-10290/2020\|EPI_ISL_583018\|2020- | .................... | .................... | .................... | .................... | .................... | .................... | |
| L | 9633 hCoV-19/USA/CA-CZB-10292/2020\|EPI_ISL_583019\|2020- | .................... | .A.................. | .................... | .................... | .................... | .................... | |
| L | 9634 hCoV-19/USA/CA-CZB-10293/2020\|EPI_ISL_583020\|2020- | .................... | .A.................. | .................... | .................... | .................... | .................... | |
| L | 9635 hCoV-19/USA/CA-CZB-10294/2020\|EPI_ISL_583021\|2020- | .................... | .A.................. | .................... | .................... | .................... | .................... | |
| L | 9636 hCoV-19/USA/CA-CZB-10295/2020\|EPI_ISL_583022\|2020- | .................... | .A.................. | .................... | .................... | .................... | .................... | |
| L | 9637 hCoV-19/USA/CA-CZB-10296/2020\|EPI_ISL_583023\|2020- | .................... | .A.................. | .................... | .................... | .................... | .................... | |
| L | 9638 hCoV-19/USA/CA-CZB-10300/2020\|EPI_ISL_583024\|2020- | .................... | .A.................. | .................... | .................... | .................... | .................... | |
| L | 9639 hCoV-19/USA/CA-CZB-10302/2020\|EPI_ISL_583025\|2020- | .................... | .A.................. | .................... | .................... | .................... | .................... | |
| L | 9640 hCoV-19/USA/CA-CZB-10303/2020\|EPI_ISL_583026\|2020- | .................... | .A.................. | .................... | .................... | .................... | .................... | |
| L | 9641 hCoV-19/USA/CA-CZB-10310/2020\|EPI_ISL_583028\|2020- | .................... | .................... | .................... | .................... | .................... | .................... | |
| L | 9642 hCoV-19/USA/CA-CZB-10311/2020\|EPI_ISL_583029\|2020- | .................... | .A.................. | .................... | .................... | .................... | .................... | |
| L | 9643 hCoV-19/USA/CA-CZB-10946/2020\|EPI_ISL_583032\|2020- | .................... | .................... | .................... | .................... | .................... | .................... | |

L 9644 hCoV-19/USA/CA-CZB-10947/2020|EPI_ISL_583033|2020- .................... .A.................. .................... .................... .................... ....................

Group 4- Oct 2020-Nov 2020

| A | B | C | D | E | F | G | H | I | J | K | L |
|---|---|---|---|---|---|---|---|---|---|---|---|
| Database | Database ID | GISAID ID | GCTGCAATCGTGCTACAACT | TGAACTGTTGCGACTACGTG | TTCGGAAGAGACAGGTACGTT | CACACAATCGAAGCGCAGTA | GCTGGTGCTGCAGCTTATTA | AGGGTCAAGTGCACAGTCTA | mismatches? | mismatches? | primer set? |
| M | 1 | hCoV-19/Austria/CeMM0808/2020\|EPI_ISL_583710\|2020- | .................... | .................... | .................... | .................... | .................... | .................... | | | |
| M | 2 | hCoV-19/Austria/CeMM0809/2020\|EPI_ISL_583711\|2020- | .................... | .................... | .................... | .................... | .................... | .................... | | | |
| M | 3 | hCoV-19/Austria/CeMM0810/2020\|EPI_ISL_583712\|2020- | .................... | .................... | .................... | .................... | .................... | .................... | | | |
| M | 4 | hCoV-19/Austria/CeMM0811/2020\|EPI_ISL_583713\|2020- | .................... | .................... | .................... | .................... | .................... | .................... | | | |
| M | 6 | hCoV-19/Austria/CeMM0813/2020\|EPI_ISL_583715\|2020- | .................... | .................... | .................... | .................... | .................... | .................... | | | |
| M | 7 | hCoV-19/Austria/CeMM0818/2020\|EPI_ISL_583716\|2020- | .................... | .................... | .................... | .................... | .................... | ...............R.... | | | |
| M | 8 | hCoV-19/Austria/CeMM0822/2020\|EPI_ISL_583718\|2020- | .................... | .................... | .................... | .................... | .................... | .................... | | | |
| M | 9 | hCoV-19/Austria/CeMM0823/2020\|EPI_ISL_583719\|2020- | .................... | .................... | .................... | .................... | .................... | .................... | | | |
| M | 10 | hCoV-19/Austria/CeMM0824/2020\|EPI_ISL_583720\|2020- | .................... | .................... | .................... | .................... | .................... | .................... | | | |
| M | 11 | hCoV-19/Austria/CeMM0825/2020\|EPI_ISL_583721\|2020- | .................... | .................... | .................... | .................... | .................... | .................... | | | |
| M | 12 | hCoV-19/Austria/CeMM0826/2020\|EPI_ISL_583722\|2020- | .................... | .................... | .................... | .................... | .................... | .................... | | | |
| M | 13 | hCoV-19/Austria/CeMM0828/2020\|EPI_ISL_583723\|2020- | .................... | .................... | .................... | .................... | .................... | .................... | | | |
| M | 14 | hCoV-19/Austria/CeMM0829/2020\|EPI_ISL_583724\|2020- | .................... | .................... | .................... | .................... | .................... | .................... | | | |
| M | 15 | hCoV-19/Austria/CeMM0830/2020\|EPI_ISL_583725\|2020- | .................... | .................... | .................... | .................... | .................... | .................... | | | |
| M | 16 | hCoV-19/Austria/CeMM0831/2020\|EPI_ISL_583726\|2020- | .................... | .................... | .................... | .................... | .................... | .................... | | | |
| M | 17 | hCoV-19/Austria/CeMM0851/2020\|EPI_ISL_583727\|2020- | .................... | .................... | .................... | .................... | .................... | .................... | | | |
| M | 18 | hCoV-19/Austria/CeMM0853/2020\|EPI_ISL_583728\|2020- | .................... | .................... | .................... | .................... | .................... | .................... | | | |
| M | 19 | hCoV-19/Austria/CeMM0855/2020\|EPI_ISL_583729\|2020- | .................... | .................... | .................... | .................... | .................... | .................... | | | |
| M | 20 | hCoV-19/Austria/CeMM0856/2020\|EPI_ISL_583730\|2020- | .................... | .................... | .................... | .................... | .................... | .................... | | | |
| M | 21 | hCoV-19/Austria/CeMM0865/2020\|EPI_ISL_583731\|2020- | .................... | .................... | .................... | .................... | .................... | .................... | | | |
| M | 22 | hCoV-19/Austria/CeMM0867/2020\|EPI_ISL_583732\|2020- | .................... | .................... | .................... | .................... | .................... | .................... | | | |
| M | 23 | hCoV-19/Austria/CeMM0868/2020\|EPI_ISL_583733\|2020- | .................... | .................... | .................... | .................... | .................... | .................... | | | |
| M | 24 | hCoV-19/Austria/CeMM0870/2020\|EPI_ISL_583735\|2020- | .................... | .................... | .................... | .................... | .................... | .................... | | | |
| M | 25 | hCoV-19/Austria/CeMM0872/2020\|EPI_ISL_583736\|2020- | .................... | .................... | .................... | .................... | .................... | .................... | | | |
| M | 26 | hCoV-19/Austria/CeMM0874/2020\|EPI_ISL_583737\|2020- | .................... | .................... | .................... | .................... | .................... | .................... | | | |
| M | 27 | hCoV-19/Austria/CeMM0875/2020\|EPI_ISL_583738\|2020- | .................... | .................... | .................... | .................... | .................... | .................... | | | |
| M | 28 | hCoV-19/Austria/CeMM0877/2020\|EPI_ISL_583739\|2020- | .................... | .................... | .................... | .................... | .................... | .................... | | | |
| M | 29 | hCoV-19/Austria/CeMM0878/2020\|EPI_ISL_583740\|2020- | .................... | .................... | .................... | .................... | .................... | .................... | | | |
| M | 30 | hCoV-19/Austria/CeMM0885/2020\|EPI_ISL_583741\|2020- | .................... | .................... | .................... | .................... | .................... | .................... | | | |
| M | 31 | hCoV-19/Austria/CeMM0886/2020\|EPI_ISL_583742\|2020- | .................... | .................... | .................... | .................... | .................... | .................... | | | |
| M | 32 | hCoV-19/Austria/CeMM0887/2020\|EPI_ISL_583743\|2020- | .................... | .................... | .................... | .................... | .................... | .................... | | | |
| M | 34 | hCoV-19/Austria/CeMM0892/2020\|EPI_ISL_583745\|2020- | .................... | .................... | .................... | .................... | .................... | .................... | | | |
| M | 35 | hCoV-19/Austria/CeMM0893/2020\|EPI_ISL_583746\|2020- | .................... | .................... | .................... | .................... | .................... | .................... | | | |
| M | 36 | hCoV-19/Austria/CeMM0894/2020\|EPI_ISL_583747\|2020- | .................... | .................... | .................... | .................... | .................... | .................... | | | |
| M | 37 | hCoV-19/Austria/CeMM0895/2020\|EPI_ISL_583748\|2020- | .................... | .................... | .................... | .................... | .................... | .................... | | | |
| M | 38 | hCoV-19/Austria/CeMM0896/2020\|EPI_ISL_583749\|2020- | .................... | .................... | .................... | .................... | .................... | .................... | | | |
| M | 39 | hCoV-19/Austria/CeMM0897/2020\|EPI_ISL_583750\|2020- | .................... | .................... | .................... | .................... | .................... | .................... | | | |
| M | 40 | hCoV-19/Austria/CeMM0898/2020\|EPI_ISL_583751\|2020- | .................... | .................... | .................... | .................... | .................... | .................... | | | |
| M | 41 | hCoV-19/Austria/CeMM0899/2020\|EPI_ISL_583752\|2020- | .................... | .................... | .................... | .................... | .................... | .................... | | | |
| M | 42 | hCoV-19/Austria/CeMM0900/2020\|EPI_ISL_583753\|2020- | .................... | .................... | .................... | .................... | .................... | .................... | | | |
| M | 43 | hCoV-19/Austria/CeMM0901/2020\|EPI_ISL_583754\|2020- | .................... | .................... | .................... | .................... | .................... | .................... | | | |
| M | 44 | hCoV-19/Austria/CeMM0902/2020\|EPI_ISL_583755\|2020- | .................... | .................... | .................... | .................... | .................... | .................... | | | |
| M | 46 | hCoV-19/Austria/CeMM0904/2020\|EPI_ISL_583757\|2020- | .................... | .................... | .................... | .................... | .................... | .................... | | | |
| M | 47 | hCoV-19/Austria/CeMM0905/2020\|EPI_ISL_583758\|2020- | .................... | .................... | .................... | .................... | .................... | .................... | | | |
| M | 48 | hCoV-19/Austria/CeMM0906/2020\|EPI_ISL_583759\|2020- | .................... | .................... | .................... | .................... | .................... | .................... | | | |
| M | 49 | hCoV-19/Austria/CeMM0907/2020\|EPI_ISL_583760\|2020- | .................... | .................... | .................... | .................... | .................... | .................... | | | |
| M | 50 | hCoV-19/Austria/CeMM0908/2020\|EPI_ISL_583761\|2020- | .................... | .................... | .................... | .................... | .................... | .................... | | | |
| M | 51 | hCoV-19/Austria/CeMM0909/2020\|EPI_ISL_583762\|2020- | .................... | .................... | .................... | .................... | .................... | .................... | | | |
| M | 52 | hCoV-19/Austria/CeMM0910/2020\|EPI_ISL_583763\|2020- | .................... | .................... | .................... | .................... | .................... | .................... | | | |
| M | 53 | hCoV-19/Austria/CeMM0912/2020\|EPI_ISL_583764\|2020- | .................... | .................... | .................... | .................... | .................... | .................... | | | |
| M | 54 | hCoV-19/Austria/CeMM0914/2020\|EPI_ISL_583766\|2020- | .................... | .................... | .................... | .................... | .................... | .................... | | | |
| M | 55 | hCoV-19/Austria/CeMM0915/2020\|EPI_ISL_583767\|2020- | .................... | .................... | .................... | .................... | .................... | .................... | | | |
| M | 56 | hCoV-19/Austria/CeMM0916/2020\|EPI_ISL_583768\|2020- | .................... | .................... | .................... | .................... | .................... | .................... | | | |
| M | 57 | hCoV-19/Austria/CeMM0917/2020\|EPI_ISL_583769\|2020- | .................... | .................... | .................... | .................... | .................... | .................... | | | |
| M | 58 | hCoV-19/Austria/CeMM0918/2020\|EPI_ISL_583770\|2020- | .................... | .................... | .................... | .................... | .................... | .................... | | | |
| M | 59 | hCoV-19/Austria/CeMM0919/2020\|EPI_ISL_583771\|2020- | .................... | .................... | .................... | .................... | .................... | .................... | | | |
| M | 60 | hCoV-19/Austria/CeMM0920/2020\|EPI_ISL_583772\|2020- | .................... | .................... | .................... | .................... | .................... | .................... | | | |
| M | 61 | hCoV-19/Austria/CeMM0921/2020\|EPI_ISL_583773\|2020- | .................... | .................... | .................... | .................... | .................... | .................... | | | |
| M | 62 | hCoV-19/Austria/CeMM0922/2020\|EPI_ISL_583774\|2020- | .................... | .................... | .................... | .................... | .................... | .................... | | | |
| M | 63 | hCoV-19/Austria/CeMM0923/2020\|EPI_ISL_583775\|2020- | .................... | .................... | .................... | .................... | .................... | .................... | | | |
| M | 64 | hCoV-19/Austria/CeMM0924/2020\|EPI_ISL_583776\|2020- | .................... | .................... | .................... | .................... | .................... | .................... | | | |
| M | 65 | hCoV-19/Austria/CeMM0925/2020\|EPI_ISL_583777\|2020- | .................... | .................... | .................... | .................... | .................... | .................... | | | |
| M | 66 | hCoV-19/Austria/CeMM0926/2020\|EPI_ISL_583778\|2020- | .................... | .................... | .................... | .................... | .................... | .................... | | | |
| M | 67 | hCoV-19/Austria/CeMM0927/2020\|EPI_ISL_583779\|2020- | .................... | .................... | .................... | .................... | .................... | .................... | | | |
| M | 68 | hCoV-19/Austria/CeMM0928/2020\|EPI_ISL_583780\|2020- | .................... | .................... | .................... | .................... | .................... | .................... | | | |
| M | 69 | hCoV-19/Austria/CeMM0929/2020\|EPI_ISL_583781\|2020- | .................... | .................... | .................... | .................... | .................... | .................... | | | |
| M | 70 | hCoV-19/Austria/CeMM0930/2020\|EPI_ISL_583782\|2020- | .................... | .................... | .................... | .................... | .................... | .................... | | | |
| M | 71 | hCoV-19/Austria/CeMM0931/2020\|EPI_ISL_583783\|2020- | .................... | .................... | .................... | .................... | .................... | .................... | | | |
| M | 72 | hCoV-19/Austria/CeMM0932/2020\|EPI_ISL_583784\|2020- | .................... | .................... | .................... | .................... | .................... | .................... | | | |

| | | | | | | | | |
|---|---|---|---|---|---|---|---|---|
| M | 73 | hCoV-19/Austria/CeMM0933/2020\|EPI_ISL_583785\|2020- | ···················· | ···················· | ···················· | ···················· | ···················· | ···················· |
| M | 74 | hCoV-19/Austria/CeMM0934/2020\|EPI_ISL_583786\|2020- | ···················· | ···················· | ···················· | ···················· | ···················· | ···················· |
| M | 75 | hCoV-19/Austria/CeMM0935/2020\|EPI_ISL_583787\|2020- | ···················· | ···················· | ···················· | ···················· | ···················· | ···················· |
| M | 76 | hCoV-19/Austria/CeMM0936/2020\|EPI_ISL_583788\|2020- | ···················· | ···················· | ···················· | ···················· | ···················· | ···················· |
| M | 77 | hCoV-19/Austria/CeMM0937/2020\|EPI_ISL_583789\|2020- | ···················· | ···················· | ···················· | ···················· | ···················· | ···················· |
| M | 78 | hCoV-19/Austria/CeMM0938/2020\|EPI_ISL_583790\|2020- | ···················· | ···················· | ···················· | ···················· | ···················· | ···················· |
| M | 79 | hCoV-19/Austria/CeMM0939/2020\|EPI_ISL_583791\|2020- | ···················· | ···················· | ···················· | ···················· | ···················· | ···················· |
| M | 80 | hCoV-19/Austria/CeMM0940/2020\|EPI_ISL_583792\|2020- | ···················· | ···················· | ···················· | ···················· | ···················· | ···················· |
| M | 81 | hCoV-19/Austria/CeMM0941/2020\|EPI_ISL_583793\|2020- | ···················· | ···················· | ···················· | ···················· | ···················· | ···················· |
| M | 82 | hCoV-19/England/ALDP-A2F863/2020\|EPI_ISL_587237\|20 | ···················· | ···················· | ···················· | ···················· | ···················· | ···················· |
| M | 83 | hCoV-19/England/ALDP-A2F1D1/2020\|EPI_ISL_587239\|20 | ···················· | ···················· | ···················· | ···················· | ···················· | ···················· |
| M | 84 | hCoV-19/England/ALDP-A2F414/2020\|EPI_ISL_587241\|20 | ···················· | ···················· | ···················· | ···················· | ···················· | ···················· |
| M | 85 | hCoV-19/England/ALDP-A2ED1A/2020\|EPI_ISL_587242\|20 | ···················· | ·R·················· | ···················· | ···················· | ···················· | ···R················ |
| M | 86 | hCoV-19/England/ALDP-A2E6A6/2020\|EPI_ISL_587243\|20 | ···················· | ···················· | ···················· | ···················· | ···················· | ···················· |
| M | 87 | hCoV-19/England/ALDP-A2F7EE/2020\|EPI_ISL_587244\|20 | ···················· | ···················· | ···················· | ···················· | ···················· | ···················· |
| M | 88 | hCoV-19/England/ALDP-A2F274/2020\|EPI_ISL_587245\|20 | ···················· | ···················· | ···················· | ···················· | ···················· | ···················· |
| M | 89 | hCoV-19/England/ALDP-A2E8DD/2020\|EPI_ISL_587246\|20 | ···················· | ···················· | ···················· | ···················· | ···················· | ···················· |
| M | 90 | hCoV-19/England/ALDP-A2F2B0/2020\|EPI_ISL_587247\|20 | ···················· | ···················· | ···················· | ···················· | ···················· | ···················· |
| M | 91 | hCoV-19/England/ALDP-A2EF8D/2020\|EPI_ISL_587248\|20 | ···················· | ···················· | ···················· | ···················· | ······T············· | ···················· |
| M | 92 | hCoV-19/England/ALDP-A2EBD4/2020\|EPI_ISL_587249\|20 | ···················· | ···················· | ···················· | ···················· | ···················· | ···················· |
| M | 93 | hCoV-19/England/ALDP-A2FD28/2020\|EPI_ISL_587252\|20 | ···················· | ·A·················· | ···················· | ···················· | ···················· | ···················· |
| M | 94 | hCoV-19/England/ALDP-A2F283/2020\|EPI_ISL_587253\|20 | ···················· | ···················· | ···················· | ···················· | ···················· | ···················· |
| M | 95 | hCoV-19/England/ALDP-A2FCA3/2020\|EPI_ISL_587254\|20 | ···················· | ···················· | ···················· | ···················· | ···················· | ···················· |
| M | 96 | hCoV-19/England/ALDP-A2F06B/2020\|EPI_ISL_587259\|20 | ···················· | ···················· | ···················· | ···················· | ···················· | ···················· |
| M | 97 | hCoV-19/England/ALDP-A2FC94/2020\|EPI_ISL_587260\|20 | ···················· | ···················· | ···················· | ···················· | ···················· | ···················· |
| M | 98 | hCoV-19/England/ALDP-A2EF32/2020\|EPI_ISL_587261\|20 | ···················· | ···················· | ···················· | ···················· | ···················· | ···················· |
| M | 99 | hCoV-19/England/ALDP-A2F238/2020\|EPI_ISL_587262\|20 | ···················· | ···················· | ···················· | ···················· | ···················· | ···················· |
| M | 100 | hCoV-19/England/ALDP-A2EC3B/2020\|EPI_ISL_587263\|20 | ···················· | ···················· | ···················· | ···················· | ···················· | ···················· |
| M | 101 | hCoV-19/England/ALDP-A2F089/2020\|EPI_ISL_587264\|20 | ···················· | ···················· | ···················· | ···················· | ···················· | ···················· |
| M | 102 | hCoV-19/England/ALDP-A2F60F/2020\|EPI_ISL_587266\|20 | ···················· | ···················· | ···················· | ···················· | ···················· | ···················· |
| M | 103 | hCoV-19/England/ALDP-A2E71C/2020\|EPI_ISL_587267\|20 | ···················· | ···················· | ···················· | ···················· | ···················· | ···················· |
| M | 104 | hCoV-19/England/ALDP-A2F818/2020\|EPI_ISL_587268\|20 | ···················· | ···················· | ···················· | ···················· | ···················· | ···················· |
| M | 105 | hCoV-19/England/ALDP-A2F5E4/2020\|EPI_ISL_587269\|20 | ···················· | ···················· | ···················· | ···················· | ···················· | ···················· |
| M | 106 | hCoV-19/England/ALDP-A2F0C5/2020\|EPI_ISL_587270\|20 | ···················· | ···················· | ···················· | ···················· | ···················· | ···················· |
| M | 107 | hCoV-19/England/ALDP-A2EF14/2020\|EPI_ISL_587271\|20 | ···················· | ···················· | ···················· | ···················· | ···················· | ···················· |
| M | 108 | hCoV-19/England/ALDP-A2F49C/2020\|EPI_ISL_587272\|20 | ···················· | ···················· | ···················· | ···················· | ···················· | ···················· |
| M | 109 | hCoV-19/England/ALDP-A2F942/2020\|EPI_ISL_587273\|20 | ···················· | ···················· | ···················· | ···················· | ···················· | ···················· |
| M | 110 | hCoV-19/England/ALDP-A2EECC/2020\|EPI_ISL_587274\|20 | ···················· | ···················· | ···················· | ···················· | ···················· | ···················· |
| M | 111 | hCoV-19/England/ALDP-A2FBC4/2020\|EPI_ISL_587275\|20 | ···················· | ···················· | ···················· | ···················· | ···················· | ···················· |
| M | 112 | hCoV-19/England/ALDP-A2F308/2020\|EPI_ISL_587276\|20 | ···················· | ·A·················· | ···················· | ···················· | ···················· | ···A················ |
| M | 113 | hCoV-19/England/ALDP-A2E749/2020\|EPI_ISL_587277\|20 | ···················· | ···················· | ···················· | ···················· | ···················· | ···················· |
| M | 114 | hCoV-19/England/ALDP-A2EBB6/2020\|EPI_ISL_587278\|20 | ···················· | ···················· | ···················· | ···················· | ···················· | ···················· |
| M | 115 | hCoV-19/England/ALDP-A2E9AD/2020\|EPI_ISL_587279\|20 | ···················· | ···················· | ···················· | ···················· | ···················· | ···················· |
| M | 116 | hCoV-19/England/QEUH-9F3984/2020\|EPI_ISL_588568\|20 | ···················· | ···················· | ···················· | ···················· | ···················· | ···················· |
| M | 117 | hCoV-19/England/QEUH-9F36E7/2020\|EPI_ISL_588569\|20 | ···················· | ···················· | ···················· | ···················· | ···················· | ···················· |
| M | 118 | hCoV-19/England/QEUH-9F373F/2020\|EPI_ISL_588570\|20 | ···················· | ···················· | ···················· | ···················· | ···················· | ···················· |
| M | 119 | hCoV-19/England/QEUH-9F38C3/2020\|EPI_ISL_588571\|20 | ···················· | ···················· | ···················· | ···················· | ···················· | ···················· |
| M | 120 | hCoV-19/England/QEUH-9F39A2/2020\|EPI_ISL_588573\|20 | ···················· | ···················· | ···················· | ···················· | ···················· | ···················· |
| M | 121 | hCoV-19/England/QEUH-9F34CF/2020\|EPI_ISL_588574\|20 | ···················· | ···················· | ···················· | ···················· | ···················· | ···················· |
| M | 122 | hCoV-19/England/QEUH-9F3B9D/2020\|EPI_ISL_588576\|20 | ···················· | ···················· | ···················· | ···················· | ···················· | ···················· |
| M | 123 | hCoV-19/England/QEUH-9F334A/2020\|EPI_ISL_588577\|20 | ···················· | ···················· | ···················· | ···················· | ······T············· | ···················· |
| M | 124 | hCoV-19/England/QEUH-9F376C/2020\|EPI_ISL_588578\|20 | ···················· | ···················· | ···················· | ···················· | ···················· | ···················· |
| M | 125 | hCoV-19/England/QEUH-9F3B51/2020\|EPI_ISL_588579\|20 | ···················· | ···················· | ···················· | ···················· | ···················· | ···················· |
| M | 126 | hCoV-19/England/QEUH-9F4051/2020\|EPI_ISL_588582\|20 | ···················· | ···················· | ···················· | ···················· | ···················· | ···················· |
| M | 127 | hCoV-19/England/QEUH-9F432B/2020\|EPI_ISL_588584\|20 | ···················· | ···················· | ···················· | ···················· | ···················· | ···················· |
| M | 128 | hCoV-19/England/QEUH-9F430D/2020\|EPI_ISL_588590\|20 | ···················· | ···················· | ···················· | ···················· | ···················· | ···················· |
| M | 129 | hCoV-19/England/QEUH-9F35CC/2020\|EPI_ISL_588591\|20 | ···················· | ···················· | ···················· | ···················· | ···················· | ···················· |
| M | 130 | hCoV-19/England/QEUH-9F421F/2020\|EPI_ISL_588592\|20 | ···················· | ···················· | ···················· | ···················· | ···················· | ···················· |
| M | 131 | hCoV-19/England/QEUH-9F3702/2020\|EPI_ISL_588593\|20 | ···················· | ···················· | ···················· | ···················· | ···················· | ···················· |
| M | 132 | hCoV-19/England/QEUH-9F39B1/2020\|EPI_ISL_588594\|20 | ···················· | ·A·················· | ···················· | ···················· | ···················· | ···················· |
| M | 133 | hCoV-19/England/QEUH-9F3F64/2020\|EPI_ISL_588595\|20 | ···················· | ···A················ | ···················· | ···················· | ···················· | ···················· |
| M | 134 | hCoV-19/England/QEUH-9F3E2B/2020\|EPI_ISL_588596\|20 | ···················· | ···················· | ···················· | ···················· | ···················· | ···················· |
| M | 135 | hCoV-19/England/QEUH-9F2FDE/2020\|EPI_ISL_588597\|20 | ···················· | ···················· | ···················· | ···················· | ···················· | ···················· |
| M | 136 | hCoV-19/England/QEUH-9F5542/2020\|EPI_ISL_588598\|20 | ···················· | ···················· | ···················· | ···················· | ···················· | ···················· |
| M | 137 | hCoV-19/England/QEUH-9F3456/2020\|EPI_ISL_588599\|20 | ···················· | ···················· | ···················· | ···················· | ···················· | ···················· |
| M | 138 | hCoV-19/England/QEUH-9D06E0/2020\|EPI_ISL_589899\|20 | ···················· | ···················· | ···················· | ···················· | ···················· | ···················· |
| M | 139 | hCoV-19/Norway/3328/2020\|EPI_ISL_590880\|2020-09-04 | ···················· | ···················· | ···················· | ···················· | ···················· | ···················· |
| M | 140 | hCoV-19/Norway/3361/2020\|EPI_ISL_590881\|2020-09-08 | ···················· | ···················· | ···················· | ···················· | ···················· | ···················· |
| M | 141 | hCoV-19/Norway/3365/2020\|EPI_ISL_590882\|2020-09-03 | ···················· | ···················· | ···················· | ···················· | ···················· | ···················· |
| M | 142 | hCoV-19/Norway/3367/2020\|EPI_ISL_590883\|2020-09-09 | ···················· | ···················· | ···················· | ···················· | ···················· | ···················· |
| M | 143 | hCoV-19/Norway/3368/2020\|EPI_ISL_590884\|2020-09-09 | ···················· | ···················· | ···················· | ···················· | ···················· | ···················· |
| M | 144 | hCoV-19/Norway/3376/2020\|EPI_ISL_590885\|2020-08-26 | ···················· | ···················· | ···················· | ···················· | ···················· | ···················· |
| M | 145 | hCoV-19/Norway/3378/2020\|EPI_ISL_590886\|2020-08-27 | ···················· | ···················· | ···················· | ···················· | ···················· | ···················· |
| M | 146 | hCoV-19/Norway/3381/2020\|EPI_ISL_590887\|2020-09-11 | ···················· | ···················· | ···················· | ···················· | ···················· | ···················· |
| M | 147 | hCoV-19/Norway/3382/2020\|EPI_ISL_590888\|2020-09-13 | ···················· | ···················· | ···················· | ···················· | ···················· | ···················· |
| M | 148 | hCoV-19/Norway/3401/2020\|EPI_ISL_590889\|2020-09-10 | ···················· | ···················· | ···················· | ···················· | ···················· | ···················· |
| M | 149 | hCoV-19/Norway/3422/2020\|EPI_ISL_590893\|2020-09-13 | ···················· | ···················· | ···················· | ···················· | ···················· | ···················· |
| M | 150 | hCoV-19/Norway/3428/2020\|EPI_ISL_590894\|2020-09-09 | ···················· | ···················· | ···················· | ···················· | ···················· | ···················· |
| M | 151 | hCoV-19/Norway/3429/2020\|EPI_ISL_590895\|2020-09-09 | ···················· | ···················· | ···················· | ···················· | ···················· | ···················· |
| M | 152 | hCoV-19/Norway/3431/2020\|EPI_ISL_590896\|2020-09-10 | ···················· | ···················· | ···················· | ···················· | ···················· | ···················· |

M 155 hCoV-19/Australia/VIC15012/2020|EPI_ISL_593102|202 .................... .................... ..................... .................... .................... ....................
M 156 hCoV-19/Australia/VIC15013/2020|EPI_ISL_593103|202 .................... .................... ..................... .................... .................... ....................
M 158 hCoV-19/Australia/VIC15018/2020|EPI_ISL_593105|202 .................... .................... ..................... .................... .................... ....................
M 160 hCoV-19/Australia/VIC15021/2020|EPI_ISL_593107|202 .................... .................... ..................... .................... .................... ....................
M 161 hCoV-19/Australia/VIC15023/2020|EPI_ISL_593108|202 .................... .................... ..................... .................... .................... ....................
M 162 hCoV-19/Australia/VIC15026/2020|EPI_ISL_593109|202 .................... .................... ..................... .................... .................... ....................
M 163 hCoV-19/Australia/VIC15027/2020|EPI_ISL_593110|202 .................... .................... ..................... .................... .................... ....................
M 164 hCoV-19/USA/UT-UPHL-201001538/2020|EPI_ISL_594401| .................... .................... ..................... .................... .................... ....................
M 165 hCoV-19/USA/UT-UPHL-201006663/2020|EPI_ISL_594402| .................... .................... ..................... .................... .................... ....................
M 166 hCoV-19/USA/UT-UPHL-201021091/2020|EPI_ISL_594404| .................... .................... ..................... .................... .................... ....................
M 167 hCoV-19/USA/UT-UPHL-201007083/2020|EPI_ISL_594406| .................... .................... ..................... .................... .................... ....................
M 168 hCoV-19/USA/UT-UPHL-201013682/2020|EPI_ISL_594407| .................... .................... ..................... .................... .................... ....................
M 169 hCoV-19/USA/UT-UPHL-201023603/2020|EPI_ISL_594409| .................... .................... ..................... .................... .................... ....................
M 170 hCoV-19/USA/UT-UPHL-201024883/2020|EPI_ISL_594410| .................... .................... ..................... .................... .................... ....................
M 171 hCoV-19/USA/UT-UPHL-201024324/2020|EPI_ISL_594411| .................... .................... ..................... .................... .................... ....................
M 172 hCoV-19/USA/UT-UPHL-201022144/2020|EPI_ISL_594412| .................... .................... ..................... .................... .................... ....................
M 173 hCoV-19/USA/UT-UPHL-201017152/2020|EPI_ISL_594413| .................... .................... ..................... .................... .................... ....................
M 174 hCoV-19/USA/UT-UPHL-201004144/2020|EPI_ISL_594414| .................... .................... ..................... .................... .................... ....................
M 175 hCoV-19/USA/UT-UPHL-201001617/2020|EPI_ISL_594415| .................... .................... ..................... .................... .................... ....................
M 176 hCoV-19/USA/UT-UPHL-201016827/2020|EPI_ISL_594416| .................... .................... ..................... .................... .................... ....................
M 177 hCoV-19/USA/UT-UPHL-201031990/2020|EPI_ISL_594417| .................... .................... ..................... .................... .................... ....................
M 178 hCoV-19/USA/UT-UPHL-201005122/2020|EPI_ISL_594418| .................... .................... ..................... .................... .................... ....................
M 179 hCoV-19/USA/UT-UPHL-201004959/2020|EPI_ISL_594420| .................... .................... ..................... .................... .................... ....................
M 180 hCoV-19/USA/UT-UPHL-201010841/2020|EPI_ISL_594421| .................... .................... ..................... .................... .................... ....................
M 181 hCoV-19/USA/UT-UPHL-201008164/2020|EPI_ISL_594422| .................... .................... ..................... .................... .................... ....................
M 182 hCoV-19/USA/UT-UPHL-201021691/2020|EPI_ISL_594423| .................... .................... ..................... .................... .................... ....................
M 183 hCoV-19/USA/UT-UPHL-201016485/2020|EPI_ISL_594425| .................... .................... ..................... .................... .................... ....................
M 184 hCoV-19/USA/UT-UPHL-201013642/2020|EPI_ISL_594427| .................... .................... ..................... .................... .................... ....................
M 185 hCoV-19/USA/UT-UPHL-201002768/2020|EPI_ISL_594428| .................... .................... ..................... .................... .................... ....................
M 186 hCoV-19/USA/UT-UPHL-201002601/2020|EPI_ISL_594429| .................... .................... ..................... .................... .................... ....................
M 187 hCoV-19/USA/UT-UPHL-201012002/2020|EPI_ISL_594430| .................... .................... ..................... .................... .................... ....................
M 188 hCoV-19/USA/UT-UPHL-201010041/2020|EPI_ISL_594432| .................... .................... ..................... .................... .................... ....................
M 189 hCoV-19/England/QEUH-9F774A/2020|EPI_ISL_595700|20 .................... .................... ..................... .................... .................... ....................
M 190 hCoV-19/Scotland/QEUH-9F77B3/2020|EPI_ISL_595701|2 .................... .................... ..................... .................... .................... ....................
M 191 hCoV-19/Scotland/QEUH-9F77C2/2020|EPI_ISL_595702|2 .................... .................... ..................... .................... .................... ....................
M 192 hCoV-19/England/QEUH-9F77D1/2020|EPI_ISL_595703|20 .................... .................... ..................... .................... .................... ....................
M 193 hCoV-19/England/QEUH-9F77E0/2020|EPI_ISL_595704|20 .................... .................... ..................... .................... .................... ....................
M 194 hCoV-19/England/QEUH-9F780B/2020|EPI_ISL_595705|20 .................... .................... ..................... .................... .................... ....................
M 195 hCoV-19/England/QEUH-9F781A/2020|EPI_ISL_595706|20 .................... .................... ..................... .................... .................... ....................
M 196 hCoV-19/England/QEUH-9F7829/2020|EPI_ISL_595707|20 .................... .................... ..................... .................... .................... ....................
M 197 hCoV-19/England/QEUH-9F7838/2020|EPI_ISL_595708|20 .................... .................... ..................... .................... .................... ....................
M 198 hCoV-19/England/QEUH-9F7883/2020|EPI_ISL_595709|20 .................... .................... ..................... .................... .................... ....................
M 199 hCoV-19/England/QEUH-9F78B0/2020|EPI_ISL_595710|20 .................... .................... ..................... .................... .................... ....................
M 200 hCoV-19/England/QEUH-9F78FC/2020|EPI_ISL_595711|20 .................... .................... ..................... .................... .................... ....................
M 201 hCoV-19/England/QEUH-9F7926/2020|EPI_ISL_595712|20 .................... .................... ..................... .................... .................... ....................
M 202 hCoV-19/England/QEUH-9F7971/2020|EPI_ISL_595713|20 .................... .................... ..................... .................... .................... ....................
M 203 hCoV-19/England/QEUH-9F7980/2020|EPI_ISL_595714|20 .................... .................... ..................... .................... .................... ....................
M 204 hCoV-19/England/QEUH-9F79DB/2020|EPI_ISL_595715|20 .................... .................... ..................... .................... .................... ....................
M 205 hCoV-19/England/QEUH-9F79EA/2020|EPI_ISL_595716|20 .................... .................... ..................... .................... .................... ....................
M 206 hCoV-19/England/QEUH-9F79F9/2020|EPI_ISL_595717|20 .................... .................... ..................... .................... .................... ....................
M 207 hCoV-19/England/QEUH-9F7A14/2020|EPI_ISL_595718|20 .................... .................... ..................... .................... .................... ....................
M 208 hCoV-19/England/QEUH-9F7E72/2020|EPI_ISL_595719|20 .................... .................... ..................... .................... .................... ....................
M 209 hCoV-19/England/QEUH-9F7EBE/2020|EPI_ISL_595720|20 .................... .................... ..................... .................... .................... ....................
M 210 hCoV-19/England/QEUH-9F7EEB/2020|EPI_ISL_595721|20 .................... .................... ..................... .................... .................... ....................
M 211 hCoV-19/Scotland/QEUH-9F7EFA/2020|EPI_ISL_595722|2 .................... .................... ..................... .................... .................... ....................
M 212 hCoV-19/England/QEUH-9F7F15/2020|EPI_ISL_595723|20 .................... .................... ..................... .................... .................... ....................
M 213 hCoV-19/England/QEUH-9F7F24/2020|EPI_ISL_595724|20 .................... .................... ..................... .................... .................... ....................
M 214 hCoV-19/England/QEUH-9F8099/2020|EPI_ISL_595725|20 .................... .................... ..................... .................... .................... ....................
M 215 hCoV-19/England/QEUH-9F80A8/2020|EPI_ISL_595726|20 .................... .................... ..................... .................... .................... ....................
M 216 hCoV-19/England/QEUH-9F80B7/2020|EPI_ISL_595727|20 .................... .................... ..................... .................... .................... ....................
M 217 hCoV-19/England/QEUH-9F80E4/2020|EPI_ISL_595729|20 .................... .................... ..................... .................... .................... ....................
M 218 hCoV-19/England/QEUH-9F810F/2020|EPI_ISL_595731|20 .................... .................... ..................... .................... .................... ....................
M 219 hCoV-19/England/QEUH-9F812D/2020|EPI_ISL_595732|20 .................... .................... ..................... .................... .................... ....................
M 220 hCoV-19/England/QEUH-9F8381/2020|EPI_ISL_595733|20 .................... .................... ..................... .................... .................... ....................
M 221 hCoV-19/England/QEUH-9F8390/2020|EPI_ISL_595734|20 .................... .................... ..................... .................... .................... ....................
M 222 hCoV-19/England/QEUH-9F83BE/2020|EPI_ISL_595735|20 .................... .................... ..................... .................... .................... ....................
M 223 hCoV-19/England/QEUH-9F83EB/2020|EPI_ISL_595736|20 .................... .................... ..................... .................... .................... ....................
M 224 hCoV-19/England/QEUH-9F83FA/2020|EPI_ISL_595737|20 .................... .................... ..................... .................... .................... ....................
M 225 hCoV-19/England/QEUH-9F8415/2020|EPI_ISL_595738|20 .................... .................... ..................... .................... .................... ....................
M 226 hCoV-19/England/QEUH-9F92A1/2020|EPI_ISL_595739|20 .................... .................... ..................... .................... .................... ....................
M 227 hCoV-19/England/QEUH-9F998E/2020|EPI_ISL_595740|20 .................... .................... ..................... .................... .................... ....................
M 228 hCoV-19/England/QEUH-9F999D/2020|EPI_ISL_595741|20 .................... .A.................. ..................... .................... .................... ..A.................
M 229 hCoV-19/England/QEUH-9F99BB/2020|EPI_ISL_595742|20 .................... .................... ..................... .................... .................... ....................
M 230 hCoV-19/England/QEUH-9F99F7/2020|EPI_ISL_595743|20 .................... .................... ..................... .................... .................... ....................
M 231 hCoV-19/England/QEUH-9F9A03/2020|EPI_ISL_595744|20 .................... .................... ..................... .................... .................... ....................
M 232 hCoV-19/England/QEUH-9F9A30/2020|EPI_ISL_595745|20 .................... .A.................. ..................... .................... .................... ....................
M 233 hCoV-19/England/QEUH-9F9AB8/2020|EPI_ISL_595746|20 .................... .................... ..................... .................... .................... ....................
M 234 hCoV-19/England/QEUH-9F9AC7/2020|EPI_ISL_595747|20 .................... .................... ..................... .................... .................... ....................
M 235 hCoV-19/England/QEUH-9F9B6A/2020|EPI_ISL_595749|20 .................... .................... ..................... .................... .................... ....................
M 236 hCoV-19/England/QEUH-9F9B88/2020|EPI_ISL_595750|20 .................... .................... ..................... .................... .................... ....................

| | | | | | | | | |
|---|---|---|---|---|---|---|---|---|
| M | 237 | hCoV-19/England/QEUH-9F9B97/2020\|EPI_ISL_595751\|20 | ···················· | ···················· | ····················· | ···················· | ······T············· | ···················· |
| M | 238 | hCoV-19/England/QEUH-9F9BE2/2020\|EPI_ISL_595753\|20 | ···················· | ···················· | ····················· | ···················· | ···················· | ···················· |
| M | 239 | hCoV-19/England/QEUH-9F9C3A/2020\|EPI_ISL_595754\|20 | ···················· | ···················· | ····················· | ···················· | ···················· | ···················· |
| M | 240 | hCoV-19/England/QEUH-9F9C49/2020\|EPI_ISL_595755\|20 | ···················· | ···················· | ····················· | ···················· | ···················· | ···················· |
| M | 241 | hCoV-19/England/QEUH-9F9C76/2020\|EPI_ISL_595757\|20 | ···················· | ···················· | ····················· | ···················· | ···················· | ···················· |
| M | 242 | hCoV-19/Wales/QEUH-9F9CFE/2020\|EPI_ISL_595758\|2020 | ···················· | ···················· | ····················· | ···················· | ···················· | ···················· |
| M | 243 | hCoV-19/England/QEUH-9F9D0A/2020\|EPI_ISL_595759\|20 | ···················· | ···················· | ····················· | ···················· | ···················· | ···················· |
| M | 244 | hCoV-19/England/QEUH-9F9D19/2020\|EPI_ISL_595760\|20 | ···················· | ···················· | ····················· | ···················· | ······T············· | ···················· |
| M | 245 | hCoV-19/England/QEUH-9F9D28/2020\|EPI_ISL_595761\|20 | ···················· | ···················· | ····················· | ···················· | ···················· | ···················· |
| M | 246 | hCoV-19/England/QEUH-9F9D37/2020\|EPI_ISL_595762\|20 | ···················· | ···················· | ····················· | ···················· | ···················· | ···················· |
| M | 247 | hCoV-19/England/QEUH-9F9D46/2020\|EPI_ISL_595763\|20 | ···················· | ···················· | ····················· | ···················· | ···················· | ···················· |
| M | 248 | hCoV-19/England/QEUH-9F9D64/2020\|EPI_ISL_595764\|20 | ···················· | ···················· | ····················· | ···················· | ···················· | ···················· |
| M | 249 | hCoV-19/England/QEUH-9F9DA0/2020\|EPI_ISL_595765\|20 | ···················· | ···················· | ····················· | ···················· | ······T············· | ···················· |
| M | 250 | hCoV-19/Scotland/QEUH-9F9DCE/2020\|EPI_ISL_595766\|2 | ···················· | ···················· | ····················· | ···················· | ···················· | ···················· |
| M | 251 | hCoV-19/England/QEUH-9F9DDD/2020\|EPI_ISL_595767\|20 | ···················· | ···················· | ····················· | ···················· | ···················· | ···················· |
| M | 252 | hCoV-19/England/QEUH-9F9E34/2020\|EPI_ISL_595768\|20 | ···················· | ···················· | ···········T········· | ···················· | ···················· | ···················· |
| M | 253 | hCoV-19/England/QEUH-9F9E43/2020\|EPI_ISL_595769\|20 | ···················· | ···················· | ····················· | ···················· | ···················· | ···················· |
| M | 254 | hCoV-19/England/QEUH-9F9E61/2020\|EPI_ISL_595770\|20 | ···················· | ·A·················· | ····················· | ···················· | ···················· | ··A················· |
| M | 255 | hCoV-19/England/QEUH-9F9E70/2020\|EPI_ISL_595771\|20 | ···················· | ···················· | ····················· | ···················· | ···················· | ···················· |
| M | 256 | hCoV-19/England/QEUH-9F9EBC/2020\|EPI_ISL_595772\|20 | ···················· | ···················· | ····················· | ···················· | ···················· | ···················· |
| M | 257 | hCoV-19/Austria/CeMM0788/2020\|EPI_ISL_583700\|2020- | ···················· | ···················· | ····················· | ···················· | ···················· | ·N·················· |
| M | 258 | hCoV-19/Austria/CeMM0790/2020\|EPI_ISL_583701\|2020- | ···················· | ···················· | ····················· | ···················· | ···················· | ···················· |
| M | 259 | hCoV-19/Austria/CeMM0792/2020\|EPI_ISL_583702\|2020- | ···················· | ···················· | ····················· | ···················· | ···················· | ···················· |
| M | 260 | hCoV-19/Austria/CeMM0793/2020\|EPI_ISL_583703\|2020- | ···················· | ···················· | ····················· | ···················· | ···················· | ···················· |
| M | 261 | hCoV-19/Austria/CeMM0794/2020\|EPI_ISL_583704\|2020- | ···················· | ···················· | ····················· | ···················· | ···················· | ···················· |
| M | 262 | hCoV-19/Austria/CeMM0795/2020\|EPI_ISL_583705\|2020- | ···················· | ···················· | ····················· | ···················· | ···················· | ···················· |
| M | 263 | hCoV-19/Austria/CeMM0796/2020\|EPI_ISL_583706\|2020- | ···················· | ···················· | ····················· | ···················· | ···················· | ···················· |
| M | 264 | hCoV-19/Austria/CeMM0797/2020\|EPI_ISL_583707\|2020- | ···················· | ···················· | ····················· | ···················· | ···················· | ···················· |
| M | 265 | hCoV-19/Austria/CeMM0807/2020\|EPI_ISL_583709\|2020- | ···················· | ···················· | ····················· | ···················· | ···················· | ···················· |
| M | 266 | hCoV-19/Wales/PHWC-16F6B1/2020\|EPI_ISL_586005\|2020 | ···················· | ···················· | ····················· | ···················· | ···················· | ···················· |
| M | 267 | hCoV-19/Wales/PHWC-16F6C0/2020\|EPI_ISL_586006\|2020 | ···················· | ···················· | ····················· | ···················· | ···················· | ···················· |
| M | 269 | hCoV-19/Wales/PHWC-16F6EE/2020\|EPI_ISL_586008\|2020 | ···················· | ···················· | ····················· | ···················· | ···················· | ···················· |
| M | 270 | hCoV-19/Wales/PHWC-16F6FD/2020\|EPI_ISL_586009\|2020 | ···················· | ···················· | ····················· | ···················· | ···················· | ···················· |
| M | 271 | hCoV-19/Wales/PHWC-16F718/2020\|EPI_ISL_586011\|2020 | ···················· | ···················· | ····················· | ···················· | ···················· | ···················· |
| M | 272 | hCoV-19/Wales/PHWC-16F745/2020\|EPI_ISL_586013\|2020 | ···················· | ···················· | ····················· | ···················· | ···················· | ···················· |
| M | 273 | hCoV-19/Wales/PHWC-16F754/2020\|EPI_ISL_586014\|2020 | ···················· | ···················· | ····················· | ···················· | ···················· | ···················· |
| M | 274 | hCoV-19/Wales/PHWC-16F763/2020\|EPI_ISL_586015\|2020 | ···················· | ···················· | ····················· | ···················· | ···················· | ···················· |
| M | 275 | hCoV-19/Wales/PHWC-16F772/2020\|EPI_ISL_586016\|2020 | ···················· | ···················· | ····················· | ···················· | ···················· | ···················· |
| M | 276 | hCoV-19/Wales/PHWC-16F790/2020\|EPI_ISL_586017\|2020 | ···················· | ···················· | ····················· | ···················· | ···················· | ···················· |
| M | 278 | hCoV-19/Wales/PHWC-16F7CD/2020\|EPI_ISL_586020\|2020 | ···················· | ···················· | ····················· | ···················· | ···················· | ···················· |
| M | 279 | hCoV-19/Wales/PHWC-16F7DC/2020\|EPI_ISL_586021\|2020 | ···················· | ···················· | ····················· | ···················· | ···················· | ···················· |
| M | 280 | hCoV-19/Wales/PHWC-16F7EB/2020\|EPI_ISL_586022\|2020 | ···················· | ···················· | ····················· | ···················· | ···················· | ···················· |
| M | 281 | hCoV-19/Wales/PHWC-16F7FA/2020\|EPI_ISL_586023\|2020 | ···················· | ···················· | ····················· | ···················· | ···················· | ···················· |
| M | 282 | hCoV-19/Wales/PHWC-16F824/2020\|EPI_ISL_586025\|2020 | ···················· | ···················· | ····················· | ···················· | ···················· | ···················· |
| M | 283 | hCoV-19/Wales/PHWC-16F833/2020\|EPI_ISL_586026\|2020 | ···················· | ···················· | ····················· | ···················· | ···················· | ···················· |
| M | 284 | hCoV-19/England/ALDP-A2F47E/2020\|EPI_ISL_587250\|20 | ···················· | ···················· | ····················· | ···················· | ···················· | ···················· |
| M | 285 | hCoV-19/England/ALDP-A2FD82/2020\|EPI_ISL_587255\|20 | ···················· | ···················· | ····················· | ···················· | ···················· | ···················· |
| M | 286 | hCoV-19/England/ALDP-A2F07A/2020\|EPI_ISL_587256\|20 | ···················· | ···················· | ····················· | ···················· | ···················· | ···················· |
| M | 287 | hCoV-19/England/ALDP-A2F906/2020\|EPI_ISL_587257\|20 | ···················· | ···················· | ····················· | ···················· | ···················· | ···················· |
| M | 288 | hCoV-19/England/ALDP-A2FD73/2020\|EPI_ISL_587258\|20 | ···················· | ···················· | ····················· | ···················· | ···················· | ···················· |
| M | 289 | hCoV-19/England/ALDP-A2ED56/2020\|EPI_ISL_587280\|20 | ···················· | ···················· | ····················· | ···················· | ···················· | ···················· |
| M | 290 | hCoV-19/England/ALDP-A2F3AE/2020\|EPI_ISL_587281\|20 | ···················· | ···················· | ····················· | ···················· | ···················· | ···················· |
| M | 291 | hCoV-19/England/ALDP-A2F784/2020\|EPI_ISL_587282\|20 | ···················· | ···················· | ····················· | ···················· | ···················· | ···················· |
| M | 292 | hCoV-19/England/ALDP-A2F2ED/2020\|EPI_ISL_587283\|20 | ···················· | ···················· | ····················· | ···················· | ···················· | ···················· |
| M | 293 | hCoV-19/England/ALDP-A2E934/2020\|EPI_ISL_587284\|20 | ···················· | ···················· | ····················· | ···················· | ···················· | ···················· |
| M | 294 | hCoV-19/England/ALDP-A2FA9A/2020\|EPI_ISL_587285\|20 | ···················· | ···················· | ····················· | ···················· | ···················· | ···················· |
| M | 295 | hCoV-19/England/ALDP-A2F9BB/2020\|EPI_ISL_587286\|20 | ···················· | ···················· | ····················· | ···················· | ···················· | ···················· |
| M | 296 | hCoV-19/England/ALDP-A2F7B1/2020\|EPI_ISL_587287\|20 | ···················· | ···················· | ····················· | ···················· | ···················· | ···················· |
| M | 297 | hCoV-19/England/ALDP-A2F827/2020\|EPI_ISL_587288\|20 | ···················· | ···················· | ····················· | ···················· | ···················· | ···················· |
| M | 298 | hCoV-19/England/ALDP-A2ED83/2020\|EPI_ISL_587289\|20 | ···················· | ···················· | ····················· | ···················· | ···················· | ···················· |
| M | 299 | hCoV-19/England/ALDP-A2EE71/2020\|EPI_ISL_587290\|20 | ···················· | ···················· | ····················· | ···················· | ···················· | ···················· |
| M | 300 | hCoV-19/England/ALDP-A2FBD3/2020\|EPI_ISL_587291\|20 | ···················· | ·A·················· | ····················· | ···················· | ···················· | ··A················· |
| M | 301 | hCoV-19/England/ALDP-A2F4E7/2020\|EPI_ISL_587292\|20 | ···················· | ···················· | ····················· | ···················· | ···················· | ···················· |
| M | 302 | hCoV-19/England/ALDP-A2EB2F/2020\|EPI_ISL_587294\|20 | ···················· | ···················· | ····················· | ···················· | ···················· | ···················· |
| M | 303 | hCoV-19/England/ALDP-A2EEAE/2020\|EPI_ISL_587295\|20 | ···················· | ···················· | ····················· | ···················· | ···················· | ···················· |
| M | 304 | hCoV-19/England/ALDP-A2F5D5/2020\|EPI_ISL_587296\|20 | ···················· | ···················· | ····················· | ···················· | ···················· | ···················· |
| M | 305 | hCoV-19/England/ALDP-A2F247/2020\|EPI_ISL_587297\|20 | ···················· | ·A·················· | ····················· | ···················· | ···················· | ··A················· |
| M | 306 | hCoV-19/England/ALDP-A2E873/2020\|EPI_ISL_587298\|20 | ···················· | ···················· | ····················· | ···················· | ···················· | ···················· |
| M | 307 | hCoV-19/England/ALDP-A2E8BF/2020\|EPI_ISL_587299\|20 | ···················· | ···················· | ····················· | ···················· | ···················· | ···················· |
| M | 308 | hCoV-19/England/ALDP-A2F845/2020\|EPI_ISL_587300\|20 | ···················· | ···················· | ····················· | ···················· | ···················· | ···················· |
| M | 309 | hCoV-19/England/ALDP-A2F739/2020\|EPI_ISL_587301\|20 | ···················· | ···················· | ····················· | ···················· | ···················· | ···················· |
| M | 310 | hCoV-19/England/ALDP-A2E961/2020\|EPI_ISL_587302\|20 | ···················· | ···················· | ····················· | ···················· | ···················· | ···················· |
| M | 311 | hCoV-19/England/QEUH-A1F189/2020\|EPI_ISL_587304\|20 | ···················· | ···················· | ····················· | ···················· | ···················· | ···················· |
| M | 312 | hCoV-19/England/QEUH-A1E1A8/2020\|EPI_ISL_587305\|20 | ···················· | ···················· | ····················· | ···················· | ····T··············· | ···················· |
| M | 313 | hCoV-19/England/QEUH-A1EE65/2020\|EPI_ISL_587306\|20 | ···················· | ···················· | ····················· | ···················· | ···················· | ···················· |
| M | 314 | hCoV-19/England/QEUH-A1E4EB/2020\|EPI_ISL_587308\|20 | ···················· | ···················· | ····················· | ···················· | ···················· | ···················· |
| M | 315 | hCoV-19/England/QEUH-A1DBE7/2020\|EPI_ISL_587309\|20 | ···················· | ···················· | ····················· | ···················· | ···················· | ···················· |
| M | 316 | hCoV-19/England/QEUH-A1DC99/2020\|EPI_ISL_587310\|20 | ···················· | ···················· | ····················· | ···················· | ···················· | ···················· |
| M | 317 | hCoV-19/England/QEUH-A1E4AF/2020\|EPI_ISL_587311\|20 | ···················· | ···················· | ····················· | ···················· | ···················· | ···················· |
| M | 318 | hCoV-19/England/QEUH-A21315/2020\|EPI_ISL_587312\|20 | ···················· | ···················· | ····················· | ···················· | ···················· | ···················· |

| | | | | | | | | |
|---|---|---|---|---|---|---|---|---|
| M | 319 | hCoV-19/England/QEUH-A223E7/2020\|EPI_ISL_587313\|20 | ···················· | ·A·················· | ···················· | ···················· | ···················· | ···················· |
| M | 320 | hCoV-19/England/QEUH-A1DD0F/2020\|EPI_ISL_587337\|20 | ···················· | ···················· | ···················· | ···················· | ···················· | ···················· |
| M | 321 | hCoV-19/England/QEUH-A1E9A0/2020\|EPI_ISL_587338\|20 | ···················· | ···················· | ···················· | ···················· | ···················· | ···················· |
| M | 322 | hCoV-19/England/QEUH-A1F16B/2020\|EPI_ISL_587339\|20 | ···················· | ···················· | ···················· | ···················· | ···················· | ···················· |
| M | 323 | hCoV-19/England/QEUH-A21667/2020\|EPI_ISL_587340\|20 | ···················· | ···················· | ···················· | ···················· | ···················· | ···················· |
| M | 324 | hCoV-19/England/QEUH-A21676/2020\|EPI_ISL_587341\|20 | ···················· | ···················· | ···················· | ···················· | ···················· | ···················· |
| M | 325 | hCoV-19/England/QEUH-A1F198/2020\|EPI_ISL_587342\|20 | ···················· | ···················· | ···················· | ···················· | ···················· | ···················· |
| M | 326 | hCoV-19/England/QEUH-A1E2C3/2020\|EPI_ISL_587343\|20 | ···················· | ···················· | ···················· | ···················· | ···················· | ···················· |
| M | 327 | hCoV-19/England/QEUH-A1E560/2020\|EPI_ISL_587344\|20 | ···················· | ···················· | ···················· | ···················· | ···················· | ···················· |
| M | 328 | hCoV-19/England/QEUH-A223C9/2020\|EPI_ISL_587345\|20 | ···················· | ···················· | ···················· | ···················· | ···················· | ···················· |
| M | 329 | hCoV-19/England/QEUH-A1EE1A/2020\|EPI_ISL_587346\|20 | ···················· | ···················· | ···················· | ···················· | ···················· | ···················· |
| M | 330 | hCoV-19/England/QEUH-A1F4BD/2020\|EPI_ISL_587348\|20 | ···················· | ···················· | ···················· | ···················· | ···················· | ···················· |
| M | 331 | hCoV-19/England/QEUH-A1E82B/2020\|EPI_ISL_587349\|20 | ···················· | ···················· | ···················· | ···················· | ···················· | ···················· |
| M | 332 | hCoV-19/England/QEUH-A222EA/2020\|EPI_ISL_587350\|20 | ···················· | ·R·················· | ···················· | ···················· | ···················· | ···················· |
| M | 333 | hCoV-19/England/QEUH-A1F471/2020\|EPI_ISL_587351\|20 | ···················· | ···················· | ···················· | ···················· | ···················· | ···················· |
| M | 334 | hCoV-19/England/QEUH-A1E366/2020\|EPI_ISL_587352\|20 | ···················· | ···················· | ···················· | ···················· | ···················· | ···················· |
| M | 335 | hCoV-19/England/QEUH-A1DC4E/2020\|EPI_ISL_587353\|20 | ···················· | ···················· | ···················· | ···················· | ···················· | ···················· |
| M | 336 | hCoV-19/England/QEUH-A1EE0B/2020\|EPI_ISL_587354\|20 | ···················· | ···················· | ···················· | ···················· | ···················· | ···················· |
| M | 338 | hCoV-19/England/QEUH-A1EFF9/2020\|EPI_ISL_587356\|20 | ···················· | ···················· | ···················· | ···················· | ···················· | ···················· |
| M | 339 | hCoV-19/England/QEUH-A1DE75/2020\|EPI_ISL_587357\|20 | ···················· | ···················· | ···················· | ···················· | ···················· | ···················· |
| M | 340 | hCoV-19/England/QEUH-9F3580/2020\|EPI_ISL_588580\|20 | ···················· | ···················· | ···················· | ···················· | ···················· | ···················· |
| M | 341 | hCoV-19/England/QEUH-9F3A81/2020\|EPI_ISL_588586\|20 | ···················· | ···················· | ···················· | ···················· | ···················· | ···················· |
| M | 342 | hCoV-19/England/QEUH-9F2E59/2020\|EPI_ISL_588587\|20 | ···················· | ···················· | ···················· | ···················· | ···················· | ···················· |
| M | 343 | hCoV-19/England/QEUH-9F33E0/2020\|EPI_ISL_588588\|20 | ···················· | ···················· | ···················· | ···················· | ···················· | ···················· |
| M | 344 | hCoV-19/England/QEUH-9F5524/2020\|EPI_ISL_588589\|20 | ···················· | ···················· | ···················· | ···················· | ···················· | ···················· |
| M | 345 | hCoV-19/England/QEUH-9F385A/2020\|EPI_ISL_588602\|20 | ···················· | ·A·················· | ···················· | ···················· | ···················· | ··A················· |
| M | 346 | hCoV-19/England/QEUH-9F3210/2020\|EPI_ISL_588603\|20 | ···················· | ···················· | ···················· | ···················· | ···················· | ···················· |
| M | 347 | hCoV-19/England/QEUH-9F4033/2020\|EPI_ISL_588604\|20 | ···················· | ···················· | ···················· | ···················· | ···················· | ···················· |
| M | 348 | hCoV-19/England/QEUH-9F3517/2020\|EPI_ISL_588605\|20 | ···················· | ···················· | ···················· | ···················· | ···················· | ···················· |
| M | 349 | hCoV-19/England/QEUH-9F34FC/2020\|EPI_ISL_588606\|20 | ···················· | ···················· | ···················· | ···················· | ···················· | ···················· |
| M | 350 | hCoV-19/England/QEUH-9F31AA/2020\|EPI_ISL_588607\|20 | ···················· | ···················· | ···················· | ···················· | ···················· | ···················· |
| M | 351 | hCoV-19/England/QEUH-9F40AC/2020\|EPI_ISL_588608\|20 | ···················· | ···················· | ···················· | ···················· | ···················· | ···················· |
| M | 352 | hCoV-19/England/QEUH-9F418B/2020\|EPI_ISL_588609\|20 | ···················· | ···················· | ···················· | ···················· | ···················· | ···················· |
| M | 353 | hCoV-19/England/QEUH-9F30AD/2020\|EPI_ISL_588610\|20 | ···················· | ···················· | ···················· | ···················· | ···················· | ···················· |
| M | 354 | hCoV-19/England/QEUH-9F3EA3/2020\|EPI_ISL_588611\|20 | ···················· | ···················· | ···················· | ···················· | ···················· | ···················· |
| M | 355 | hCoV-19/England/QEUH-9F35DB/2020\|EPI_ISL_588612\|20 | ···················· | ···················· | ···················· | ···················· | ···················· | ···················· |
| M | 356 | hCoV-19/England/QEUH-9F3887/2020\|EPI_ISL_588613\|20 | ···················· | ···················· | ···················· | ···················· | ···················· | ···················· |
| M | 357 | hCoV-19/England/QEUH-9F407F/2020\|EPI_ISL_588614\|20 | ···················· | ···················· | ···················· | ···················· | ···················· | ···················· |
| M | 358 | hCoV-19/England/QEUH-9F2FCF/2020\|EPI_ISL_588615\|20 | ···················· | ···················· | ···················· | ···················· | ···················· | ···················· |
| M | 359 | hCoV-19/England/QEUH-9F3465/2020\|EPI_ISL_588616\|20 | ···················· | ···················· | ···················· | ···················· | ···················· | ···················· |
| M | 360 | hCoV-19/Wales/QEUH-9F3939/2020\|EPI_ISL_588617\|2020 | ···················· | ···················· | ···················· | ···················· | ···················· | ···················· |
| M | 361 | hCoV-19/England/QEUH-9F5490/2020\|EPI_ISL_588619\|20 | ···················· | ···················· | ···················· | ···················· | ···················· | ···················· |
| M | 362 | hCoV-19/England/QEUH-9F3CB8/2020\|EPI_ISL_588620\|20 | ···················· | ···················· | ···················· | ···················· | ···················· | ···················· |
| M | 363 | hCoV-19/England/QEUH-9F375D/2020\|EPI_ISL_588621\|20 | ···················· | ···················· | ···················· | ···················· | ···················· | ···················· |
| M | 364 | hCoV-19/England/QEUH-9F3ACD/2020\|EPI_ISL_588622\|20 | ···················· | ···················· | ···················· | ···················· | ···················· | ···················· |
| M | 365 | hCoV-19/England/QEUH-9F3562/2020\|EPI_ISL_588623\|20 | ···················· | ···················· | ···················· | ···················· | ····T··············· | ···················· |
| M | 366 | hCoV-19/England/QEUH-9F3E94/2020\|EPI_ISL_588624\|20 | ···················· | ···················· | ···················· | ···················· | ···················· | ···················· |
| M | 367 | hCoV-19/England/QEUH-9F3395/2020\|EPI_ISL_588625\|20 | ···················· | ·A·················· | ···················· | ···················· | ···················· | ···················· |
| M | 368 | hCoV-19/England/QEUH-9F559D/2020\|EPI_ISL_588626\|20 | ···················· | ···················· | ···················· | ···················· | ···················· | ···················· |
| M | 369 | hCoV-19/England/QEUH-9F3A09/2020\|EPI_ISL_588627\|20 | ···················· | ···················· | ···················· | ···················· | ···················· | ···················· |
| M | 370 | hCoV-19/England/QEUH-9F5551/2020\|EPI_ISL_588628\|20 | ···················· | ···················· | ···················· | ···················· | ···················· | ···················· |
| M | 371 | hCoV-19/England/QEUH-9F3C4F/2020\|EPI_ISL_588629\|20 | ···················· | ···················· | ···················· | ···················· | ···················· | ···················· |
| M | 372 | hCoV-19/England/QEUH-9F41F4/2020\|EPI_ISL_588630\|20 | ···················· | ···················· | ···················· | ···················· | ···················· | ···················· |
| M | 373 | hCoV-19/England/QEUH-9F3535/2020\|EPI_ISL_588631\|20 | ···················· | ···················· | ···················· | ···················· | ···················· | ···················· |
| M | 374 | hCoV-19/England/QEUH-9F4006/2020\|EPI_ISL_588632\|20 | ···················· | ···················· | ···················· | ···················· | ···················· | ···················· |
| M | 375 | hCoV-19/England/QEUH-9F4376/2020\|EPI_ISL_588633\|20 | ···················· | ···················· | ···················· | ···················· | ···················· | ···················· |
| M | 376 | hCoV-19/England/QEUH-9F381E/2020\|EPI_ISL_588634\|20 | ···················· | ···················· | ···················· | ···················· | ···················· | ···················· |
| M | 377 | hCoV-19/England/QEUH-9F3C8B/2020\|EPI_ISL_588635\|20 | ···················· | ···················· | ···················· | ···················· | ···················· | ···················· |
| M | 378 | hCoV-19/England/QEUH-9F3720/2020\|EPI_ISL_588636\|20 | ···················· | ···················· | ···················· | ···················· | ···················· | ···················· |
| M | 379 | hCoV-19/England/QEUH-9F33A4/2020\|EPI_ISL_588637\|20 | ···················· | ···················· | ···················· | ···················· | ···················· | ···················· |
| M | 380 | hCoV-19/England/QEUH-9F3E0D/2020\|EPI_ISL_588638\|20 | ···················· | ···················· | ···················· | ···················· | ···················· | ···················· |
| M | 381 | hCoV-19/England/QEUH-9F3E1C/2020\|EPI_ISL_588639\|20 | ···················· | ···················· | ···················· | ···················· | ······T············· | ···················· |
| M | 382 | hCoV-19/England/QEUH-9F3F46/2020\|EPI_ISL_588640\|20 | ···················· | ···················· | ···················· | ···················· | ···················· | ···················· |
| M | 383 | hCoV-19/England/QEUH-9F3A45/2020\|EPI_ISL_588641\|20 | ···················· | ···················· | ···················· | ···················· | ···················· | ···················· |
| M | 384 | hCoV-19/England/QEUH-9F3B15/2020\|EPI_ISL_588642\|20 | ···················· | ···················· | ···················· | ···················· | ···················· | ···················· |
| M | 385 | hCoV-19/England/QEUH-9F54BE/2020\|EPI_ISL_588643\|20 | ···················· | ···················· | ···················· | ···················· | ···················· | ···················· |
| M | 386 | hCoV-19/England/QEUH-9F3AFA/2020\|EPI_ISL_588644\|20 | ···················· | ···················· | ···················· | ···················· | ···················· | ···················· |
| M | 387 | hCoV-19/England/QEUH-9F2F74/2020\|EPI_ISL_588647\|20 | ···················· | ···················· | ···················· | ···················· | ···················· | ···················· |
| M | 388 | hCoV-19/England/QEUH-9F3ADC/2020\|EPI_ISL_588667\|20 | ···················· | ···················· | ···················· | ···················· | ···················· | ···················· |
| M | 389 | hCoV-19/England/QEUH-9F40E8/2020\|EPI_ISL_588668\|20 | ···················· | ···················· | ···················· | ···················· | ···················· | ···················· |
| M | 390 | hCoV-19/England/QEUH-9F3C03/2020\|EPI_ISL_588669\|20 | ···················· | ···················· | ···················· | ···················· | ···················· | ···················· |
| M | 391 | hCoV-19/England/QEUH-9F341A/2020\|EPI_ISL_588670\|20 | ···················· | ···················· | ···················· | ···················· | ···················· | ···················· |
| M | 392 | hCoV-19/Wales/QEUH-9F3526/2020\|EPI_ISL_588671\|2020 | ···················· | ···················· | ···················· | ···················· | ···················· | ···················· |
| M | 393 | hCoV-19/England/QEUH-9F5533/2020\|EPI_ISL_588672\|20 | ···················· | ···················· | ···················· | ···················· | ···················· | ···················· |
| M | 394 | hCoV-19/England/QEUH-9F5515/2020\|EPI_ISL_588675\|20 | ···················· | ···················· | ···················· | ···················· | ···················· | ···················· |
| M | 395 | hCoV-19/England/QEUH-9F4015/2020\|EPI_ISL_588676\|20 | ···················· | ···················· | ···················· | ···················· | ···················· | ···················· |
| M | 396 | hCoV-19/England/QEUH-9F3007/2020\|EPI_ISL_588677\|20 | ···················· | ···················· | ···················· | ···················· | ···················· | ···················· |
| M | 397 | hCoV-19/England/QEUH-9F557F/2020\|EPI_ISL_588678\|20 | ···················· | ···················· | ···················· | ···················· | ···················· | ···················· |
| M | 398 | hCoV-19/England/QEUH-9F3EB2/2020\|EPI_ISL_588679\|20 | ···················· | ···················· | ···················· | ···················· | ···················· | ···················· |
| M | 399 | hCoV-19/England/QEUH-9F323E/2020\|EPI_ISL_588680\|20 | ···················· | ·A·················· | ···················· | ···················· | ···················· | ···················· |

| | | | | | | | |
|---|---|---|---|---|---|---|---|
| M | 400 hCoV-19/England/QEUH-9F318C/2020\|EPI_ISL_588681\|20 | ···················· | ···················· | ····················· | ···················· | ···················· | ···················· |
| M | 401 hCoV-19/England/QEUH-9F3F19/2020\|EPI_ISL_588682\|20 | ···················· | ···················· | ····················· | ···················· | ···················· | ···················· |
| M | 402 hCoV-19/England/QEUH-9F31E6/2020\|EPI_ISL_588683\|20 | ···················· | ···················· | ····················· | ···················· | ···················· | ···················· |
| M | 403 hCoV-19/England/QEUH-9F3957/2020\|EPI_ISL_588684\|20 | ···················· | ···················· | ····················· | ···················· | ···················· | ···················· |
| M | 404 hCoV-19/England/QEUH-9F3E85/2020\|EPI_ISL_588685\|20 | ···················· | ···················· | ····················· | ···················· | ···················· | ···················· |
| M | 405 hCoV-19/England/QEUH-9F3544/2020\|EPI_ISL_588686\|20 | ···················· | ···················· | ····················· | ···················· | ···················· | ···················· |
| M | 406 hCoV-19/England/QEUH-9F2E3B/2020\|EPI_ISL_588687\|20 | ···················· | ···················· | ····················· | ···················· | ···················· | ···················· |
| M | 407 hCoV-19/England/QEUH-9F4024/2020\|EPI_ISL_588688\|20 | ···················· | ···················· | ····················· | ···················· | ···················· | ···················· |
| M | 408 hCoV-19/England/QEUH-9D0AC6/2020\|EPI_ISL_589932\|20 | ···················· | ···················· | ····················· | ···················· | ···················· | ···················· |
| M | 409 hCoV-19/England/QEUH-9CFC79/2020\|EPI_ISL_589933\|20 | ···················· | ···················· | ····················· | ···················· | ···················· | ···················· |
| M | 410 hCoV-19/England/QEUH-9D07B0/2020\|EPI_ISL_589935\|20 | ···················· | ·A·················· | ····················· | ···················· | ···················· | ···················· |
| M | 411 hCoV-19/England/QEUH-9CF990/2020\|EPI_ISL_589937\|20 | ···················· | ···················· | ····················· | ···················· | ···················· | ···················· |
| M | 412 hCoV-19/England/QEUH-9D0923/2020\|EPI_ISL_589938\|20 | ···················· | ···················· | ····················· | ···················· | ···················· | ···················· |
| M | 413 hCoV-19/England/QEUH-9D0C75/2020\|EPI_ISL_589939\|20 | ···················· | ···················· | ····················· | ···················· | ···················· | ···················· |
| M | 414 hCoV-19/England/QEUH-9D0B87/2020\|EPI_ISL_589940\|20 | ···················· | ···················· | ····················· | ···················· | ······T············· | ···················· |
| M | 415 hCoV-19/England/QEUH-9D0EE8/2020\|EPI_ISL_589942\|20 | ···················· | ···················· | ····················· | ···················· | ···················· | ···················· |
| M | 416 hCoV-19/England/QEUH-9D0F8B/2020\|EPI_ISL_589943\|20 | ···················· | ···················· | ····················· | ···················· | ···················· | ···················· |
| M | 417 hCoV-19/England/QEUH-9CFE19/2020\|EPI_ISL_589944\|20 | ···················· | ···················· | ····················· | ···················· | ···················· | ···················· |
| M | 418 hCoV-19/England/QEUH-9D055C/2020\|EPI_ISL_589945\|20 | ···················· | ···················· | ····················· | ···················· | ···················· | ···················· |
| M | 419 hCoV-19/England/QEUH-9D0D72/2020\|EPI_ISL_589947\|20 | ···················· | ···················· | ····················· | ···················· | ···················· | ···················· |
| M | 420 hCoV-19/England/QEUH-9D053E/2020\|EPI_ISL_589949\|20 | ···················· | ···················· | ····················· | ···················· | ···················· | ···················· |
| M | 421 hCoV-19/England/QEUH-9CFAAC/2020\|EPI_ISL_589950\|20 | ···················· | ···················· | ····················· | ···················· | ···················· | ···················· |
| M | 422 hCoV-19/England/QEUH-9D00E2/2020\|EPI_ISL_589951\|20 | ···················· | ···················· | ····················· | ···················· | ····T··············· | ···················· |
| M | 423 hCoV-19/England/QEUH-9D0AD5/2020\|EPI_ISL_589952\|20 | ···················· | ···················· | ····················· | ···················· | ···················· | ···················· |
| M | 424 hCoV-19/England/QEUH-9D071A/2020\|EPI_ISL_589953\|20 | ···················· | ···················· | ····················· | ···················· | ···················· | ···················· |
| M | 425 hCoV-19/England/QEUH-9D002E/2020\|EPI_ISL_589954\|20 | ···················· | ·A·················· | ····················· | ···················· | ···················· | ···A················ |
| M | 426 hCoV-19/England/QEUH-9D0FF4/2020\|EPI_ISL_589955\|20 | ···················· | ···················· | ····················· | ···················· | ···················· | ···················· |
| M | 427 hCoV-19/England/QEUH-9D08BD/2020\|EPI_ISL_589956\|20 | ···················· | ···················· | ····················· | ···················· | ···················· | ···················· |
| M | 428 hCoV-19/England/QEUH-9D0AF3/2020\|EPI_ISL_589957\|20 | ···················· | ···················· | ····················· | ···················· | ···················· | ···················· |
| M | 429 hCoV-19/England/QEUH-9D0E8E/2020\|EPI_ISL_589958\|20 | ···················· | ···················· | ····················· | ···················· | ······T············· | ···················· |
| M | 430 hCoV-19/England/QEUH-9D02FB/2020\|EPI_ISL_589959\|20 | ···················· | ···················· | ····················· | ···················· | ···················· | ···················· |
| M | 431 hCoV-19/England/QEUH-9CFE46/2020\|EPI_ISL_589960\|20 | ···················· | ···················· | ····················· | ···················· | ···················· | ···················· |
| M | 432 hCoV-19/England/QEUH-9D0905/2020\|EPI_ISL_589961\|20 | ···················· | ···················· | ····················· | ···················· | ···················· | ···················· |
| M | 433 hCoV-19/England/QEUH-9D06D1/2020\|EPI_ISL_589962\|20 | ···················· | ···················· | ····················· | ···················· | ···················· | ···················· |
| M | 434 hCoV-19/England/QEUH-9D0DAF/2020\|EPI_ISL_589965\|20 | ···················· | ···················· | ····················· | ···················· | ···················· | ···················· |
| M | 435 hCoV-19/England/QEUH-9D0F30/2020\|EPI_ISL_589966\|20 | ···················· | ···················· | ····················· | ···················· | ···················· | ···················· |
| M | 436 hCoV-19/England/QEUH-9D00D3/2020\|EPI_ISL_589967\|20 | ···················· | ···················· | ····················· | ···················· | ···················· | ···················· |
| M | 437 hCoV-19/England/QEUH-9CFB9A/2020\|EPI_ISL_589969\|20 | ···················· | ···················· | ····················· | ···················· | ···················· | ···················· |
| M | 438 hCoV-19/England/QEUH-9D0598/2020\|EPI_ISL_589970\|20 | ···················· | ·A·······A·········· | ····················· | ···················· | ···················· | ···················· |
| M | 439 hCoV-19/England/QEUH-9D0EBB/2020\|EPI_ISL_589971\|20 | ···················· | ···················· | ····················· | ···················· | ···················· | ···················· |
| M | 440 hCoV-19/England/QEUH-9D0DEB/2020\|EPI_ISL_589972\|20 | ···················· | ···················· | ····················· | ···················· | ···················· | ···················· |
| M | 441 hCoV-19/England/QEUH-9D01B2/2020\|EPI_ISL_589973\|20 | ···················· | ···················· | ····················· | ···················· | ···················· | ···················· |
| M | 442 hCoV-19/England/QEUH-9D0FA9/2020\|EPI_ISL_589975\|20 | ···················· | ···················· | ····················· | ···················· | ···················· | ···················· |
| M | 443 hCoV-19/England/QEUH-9D0D81/2020\|EPI_ISL_589979\|20 | ···················· | ···················· | ····················· | ···················· | ···················· | ···················· |
| M | 444 hCoV-19/England/QEUH-9D0914/2020\|EPI_ISL_589998\|20 | ···················· | ···················· | ····················· | ···················· | ···················· | ···················· |
| M | 445 hCoV-19/England/QEUH-9D0F21/2020\|EPI_ISL_589999\|20 | ···················· | ···················· | ····················· | ···················· | ···················· | ···················· |
| M | 446 hCoV-19/Norway/3405/2020\|EPI_ISL_590890\|2020-09-14 | ···················· | ···················· | ····················· | ···················· | ···················· | ···················· |
| M | 447 hCoV-19/Norway/3408/2020\|EPI_ISL_590891\|2020-09-11 | ···················· | ···················· | ····················· | ···················· | ···················· | ···················· |
| M | 448 hCoV-19/Norway/3413/2020\|EPI_ISL_590892\|2020-09-11 | ···················· | ···················· | ····················· | ···················· | ···················· | ···················· |
| M | 449 hCoV-19/Norway/3435/2020\|EPI_ISL_590897\|2020-09-13 | ···················· | ···················· | ····················· | ···················· | ···················· | ···················· |
| M | 450 hCoV-19/Norway/3362/2020\|EPI_ISL_590898\|2020-09-09 | ···················· | ···················· | ····················· | ···················· | ···················· | ···················· |
| M | 451 hCoV-19/Norway/3426/2020\|EPI_ISL_590899\|2020-09-09 | ···················· | ···················· | ····················· | ···················· | ···················· | ···················· |
| M | 452 hCoV-19/Norway/3243/2020\|EPI_ISL_590920\|2020-08-03 | ···················· | ···················· | ····················· | ···················· | ···················· | ···················· |
| M | 453 hCoV-19/Norway/3122/2020\|EPI_ISL_590921\|2020-08-07 | ···················· | ···················· | ····················· | ···················· | ···················· | ···················· |
| M | 454 hCoV-19/Norway/3129/2020\|EPI_ISL_590922\|2020-08-07 | ···················· | ···················· | ····················· | ···················· | ···················· | ···················· |
| M | 455 hCoV-19/Norway/3534/2020\|EPI_ISL_590923\|2020-09-18 | ···················· | ···················· | ····················· | ···················· | ···················· | ···················· |
| M | 456 hCoV-19/Norway/3590/2020\|EPI_ISL_590924\|2020-07-31 | ···················· | ·A·················· | ····················· | ···················· | ···················· | ···A················ |
| M | 457 hCoV-19/Norway/3593/2020\|EPI_ISL_590925\|2020-07-31 | ···················· | ·A·················· | ····················· | ···················· | ···················· | ···A················ |
| M | 458 hCoV-19/Norway/3571/2020\|EPI_ISL_590926\|2020-07-31 | ···················· | ·A·················· | ····················· | ···················· | ···················· | ···A················ |
| M | 459 hCoV-19/Norway/3532/2020\|EPI_ISL_590927\|2020-09-15 | ···················· | ···················· | ····················· | ···················· | ···················· | ···················· |
| M | 460 hCoV-19/Norway/3537/2020\|EPI_ISL_590928\|2020-09-13 | ···················· | ···················· | ····················· | ···················· | ···················· | ···················· |
| M | 461 hCoV-19/Norway/3603/2020\|EPI_ISL_590929\|2020-09-18 | ···················· | ···················· | ····················· | ···················· | ···················· | ···················· |
| M | 462 hCoV-19/Norway/3541/2020\|EPI_ISL_590930\|2020-09-10 | ···················· | ···················· | ····················· | ···················· | ···················· | ···················· |
| M | 463 hCoV-19/Norway/3625/2020\|EPI_ISL_590931\|2020-09-21 | ···················· | ···················· | ····················· | ···················· | ···················· | ···················· |
| M | 464 hCoV-19/Norway/3601/2020\|EPI_ISL_590932\|2020-09-16 | ···················· | ···················· | ····················· | ···················· | ···················· | ···················· |
| M | 465 hCoV-19/Norway/3602/2020\|EPI_ISL_590933\|2020-09-19 | ···················· | ···················· | ····················· | ···················· | ···················· | ···················· |
| M | 466 hCoV-19/Norway/3536/2020\|EPI_ISL_590934\|2020-09-05 | ···················· | ·A·················· | ····················· | ···················· | ···················· | ···A················ |
| M | 467 hCoV-19/Norway/3539/2020\|EPI_ISL_590935\|2020-09-10 | ···················· | ···················· | ····················· | ···················· | ···················· | ···················· |
| M | 468 hCoV-19/Norway/3543/2020\|EPI_ISL_590936\|2020-09-17 | ···················· | ···················· | ····················· | ···················· | ···················· | ···················· |
| M | 469 hCoV-19/Norway/3544/2020\|EPI_ISL_590937\|2020-09-18 | ···················· | ···················· | ····················· | ···················· | ···················· | ···················· |
| M | 470 hCoV-19/Norway/3578/2020\|EPI_ISL_590938\|2020-08-03 | ···················· | ·A·················· | ····················· | ···················· | ···················· | ···A················ |
| M | 471 hCoV-19/Norway/3582/2020\|EPI_ISL_590939\|2020-07-31 | ···················· | ·A·················· | ····················· | ···················· | ···················· | ···A················ |
| M | 472 hCoV-19/Norway/3585/2020\|EPI_ISL_590940\|2020-07-31 | ···················· | ·A·················· | ····················· | ···················· | ···················· | ···A················ |
| M | 473 hCoV-19/Norway/3592/2020\|EPI_ISL_590941\|2020-08-02 | ···················· | ·A·················· | ····················· | ···················· | ···················· | ···A················ |
| M | 474 hCoV-19/Norway/3594/2020\|EPI_ISL_590942\|2020-08-02 | ···················· | ·A·················· | ····················· | ···················· | ···················· | ···A················ |
| M | 475 hCoV-19/Norway/3596/2020\|EPI_ISL_590943\|2020-08-04 | ···················· | ·A·················· | ····················· | ···················· | ···················· | ···A················ |
| M | 476 hCoV-19/Norway/3598/2020\|EPI_ISL_590944\|2020-09-23 | ···················· | ·A·················· | ····················· | ···················· | ···················· | ···A················ |
| M | 477 hCoV-19/Norway/3600/2020\|EPI_ISL_590945\|2020-09-19 | ···················· | ···················· | ····················· | ···················· | ···················· | ···················· |
| M | 478 hCoV-19/Norway/3552/2020\|EPI_ISL_590946\|2020-08-06 | ···················· | ·A·················· | ····················· | ···················· | ···················· | ···A················ |
| M | 479 hCoV-19/Norway/3564/2020\|EPI_ISL_590947\|2020-07-31 | ···················· | ·A·················· | ····················· | ···················· | ···················· | ···A················ |

| | | | | | | | |
|---|---|---|---|---|---|---|---|
| M | 480 hCoV-19/Norway/3624/2020\|EPI_ISL_590948\|2020-09-21 | ···················· | ···················· | ···················· | ···················· | ···················· | ···················· |
| M | 481 hCoV-19/Norway/3572/2020\|EPI_ISL_590949\|2020-07-31 | ···················· | ·A·················· | ···················· | ···················· | ···················· | ··A················· |
| M | 482 hCoV-19/Norway/3574/2020\|EPI_ISL_590950\|2020-07-31 | ···················· | ·A·················· | ···················· | ···················· | ···················· | ··A················· |
| M | 483 hCoV-19/Norway/3977/2020\|EPI_ISL_590951\|2020-09-23 | ···················· | ···················· | ···················· | ···················· | ···················· | ···················· |
| M | 484 hCoV-19/Norway/3978/2020\|EPI_ISL_590952\|2020-09-24 | ···················· | ···················· | ···················· | ···················· | ···················· | ···················· |
| M | 485 hCoV-19/Norway/3984/2020\|EPI_ISL_590953\|2020-09-21 | ···················· | ···················· | ···················· | ···················· | ···················· | ···················· |
| M | 486 hCoV-19/Norway/3985/2020\|EPI_ISL_590954\|2020-09-24 | ···················· | ···················· | ···················· | ···················· | ···················· | ···················· |
| M | 487 hCoV-19/Norway/3986/2020\|EPI_ISL_590955\|2020-09-25 | ···················· | ···················· | ···················· | ···················· | ···················· | ···················· |
| M | 488 hCoV-19/Norway/3987/2020\|EPI_ISL_590956\|2020-09-21 | ···················· | ···················· | ···················· | ···················· | ···················· | ···················· |
| M | 489 hCoV-19/Norway/3990/2020\|EPI_ISL_590957\|2020-09-21 | ···················· | ···················· | ···················· | ···················· | ···················· | ···················· |
| M | 490 hCoV-19/Norway/3992/2020\|EPI_ISL_590958\|2020-09-21 | ···················· | ···················· | ···················· | ···················· | ···················· | ···················· |
| M | 491 hCoV-19/Norway/4003/2020\|EPI_ISL_590959\|2020-09-21 | ···················· | ···················· | ···················· | ···················· | ···················· | ···················· |
| M | 492 hCoV-19/Norway/3835/2020\|EPI_ISL_590980\|2020-09-25 | ···················· | ···················· | ···················· | ···················· | ···················· | ···················· |
| M | 493 hCoV-19/Norway/3842/2020\|EPI_ISL_590981\|2020-09-25 | ···················· | ···················· | ···················· | ···················· | ···················· | ···················· |
| M | 494 hCoV-19/Norway/3901/2020\|EPI_ISL_590982\|2020-09-14 | ···················· | ···················· | ···················· | ···················· | ···················· | ···················· |
| M | 495 hCoV-19/Norway/3906/2020\|EPI_ISL_590983\|2020-09-23 | ···················· | ···················· | ···················· | ···················· | ···················· | ···················· |
| M | 496 hCoV-19/Norway/3918/2020\|EPI_ISL_590984\|2020-09-24 | ···················· | ···················· | ···················· | ···················· | ···················· | ···················· |
| M | 497 hCoV-19/Norway/3708/2020\|EPI_ISL_590985\|2020-09-21 | ···················· | ···················· | ···················· | ···················· | ···················· | ···················· |
| M | 498 hCoV-19/Norway/3767/2020\|EPI_ISL_590986\|2020-08-07 | ···················· | ·A·················· | ···················· | ···················· | ···················· | ··A················· |
| M | 499 hCoV-19/Norway/3715/2020\|EPI_ISL_590987\|2020-09-03 | ···················· | ···················· | ···················· | ···················· | ···················· | ···················· |
| M | 500 hCoV-19/Norway/3718/2020\|EPI_ISL_590988\|2020-08-29 | ···················· | ···················· | ···················· | ···················· | ···················· | ···················· |
| M | 501 hCoV-19/Norway/3719/2020\|EPI_ISL_590989\|2020-09-03 | ···················· | ···················· | ···················· | ···················· | ···················· | ···················· |
| M | 502 hCoV-19/Norway/3725/2020\|EPI_ISL_590990\|2020-09-07 | ···················· | ···················· | ···················· | ···················· | ···················· | ···················· |
| M | 503 hCoV-19/Norway/3726/2020\|EPI_ISL_590991\|2020-09-01 | ···················· | ···················· | ···················· | ···················· | ···················· | ···················· |
| M | 504 hCoV-19/Norway/3727/2020\|EPI_ISL_590992\|2020-08-31 | ···················· | ···················· | ···················· | ···················· | ···················· | ···················· |
| M | 505 hCoV-19/Norway/3729/2020\|EPI_ISL_590993\|2020-08-31 | ···················· | ···················· | ···················· | ···················· | ···················· | ···················· |
| M | 506 hCoV-19/Norway/3731/2020\|EPI_ISL_590994\|2020-08-31 | ···················· | ···················· | ···················· | ···················· | ···················· | ···················· |
| M | 507 hCoV-19/Norway/3739/2020\|EPI_ISL_590995\|2020-09-02 | ···················· | ···················· | ···················· | ···················· | ···················· | ···················· |
| M | 508 hCoV-19/Norway/3740/2020\|EPI_ISL_590996\|2020-09-04 | ···················· | ···················· | ···················· | ···················· | ···················· | ···················· |
| M | 509 hCoV-19/Norway/3741/2020\|EPI_ISL_590997\|2020-08-30 | ···················· | ···················· | ···················· | ···················· | ···················· | ···················· |
| M | 510 hCoV-19/Norway/3743/2020\|EPI_ISL_590998\|2020-09-01 | ···················· | ···················· | ···················· | ···················· | ···················· | ···················· |
| M | 511 hCoV-19/Norway/3744/2020\|EPI_ISL_590999\|2020-09-04 | ···················· | ···················· | ···················· | ···················· | ···················· | ···················· |
| M | 512 hCoV-19/Australia/VIC15220/2020\|EPI_ISL_593199\|202 | ···················· | ···················· | ···················· | ···················· | ···················· | ···················· |
| M | 513 hCoV-19/Austria/CeMM0942/2020\|EPI_ISL_583794\|2020- | ···················· | ···················· | ···················· | ···················· | ···················· | ···················· |
| M | 514 hCoV-19/Austria/CeMM0943/2020\|EPI_ISL_583795\|2020- | ···················· | ···················· | ···················· | ···················· | ···················· | ···················· |
| M | 515 hCoV-19/Austria/CeMM0944/2020\|EPI_ISL_583796\|2020- | ···················· | ···················· | ···················· | ···················· | ···················· | ···················· |
| M | 516 hCoV-19/Austria/CeMM0945/2020\|EPI_ISL_583797\|2020- | ···················· | ···················· | ···················· | ···················· | ···················· | ···················· |
| M | 517 hCoV-19/Austria/CeMM0946/2020\|EPI_ISL_583798\|2020- | ···················· | ···················· | ···················· | ···················· | ···················· | ···················· |
| M | 518 hCoV-19/Austria/CeMM0947/2020\|EPI_ISL_583799\|2020- | ···················· | ···················· | ···················· | ···················· | ···················· | ···················· |
| M | 519 hCoV-19/Wales/PHWC-16F666/2020\|EPI_ISL_586000\|2020 | ···················· | ···················· | ···················· | ···················· | ···················· | ···················· |
| M | 520 hCoV-19/Wales/PHWC-16F684/2020\|EPI_ISL_586002\|2020 | ···················· | ···················· | ···················· | ···················· | ···················· | ···················· |
| M | 521 hCoV-19/Wales/PHWC-16F693/2020\|EPI_ISL_586003\|2020 | ···················· | ···················· | ···················· | ···················· | ···················· | ···················· |
| M | 522 hCoV-19/Wales/PHWC-16F6A2/2020\|EPI_ISL_586004\|2020 | ···················· | ···················· | ···················· | ···················· | ···················· | ···················· |
| M | 523 hCoV-19/England/QEUH-A1DD69/2020\|EPI_ISL_587314\|20 | ···················· | ···················· | ···················· | ···················· | ···················· | ···················· |
| M | 524 hCoV-19/England/QEUH-A1E858/2020\|EPI_ISL_587315\|20 | ···················· | ···················· | ···················· | ···················· | ···················· | ···················· |
| M | 525 hCoV-19/England/QEUH-A21324/2020\|EPI_ISL_587317\|20 | ···················· | ···················· | ···················· | ···················· | ···················· | ···················· |
| M | 526 hCoV-19/England/QEUH-A1DAEA/2020\|EPI_ISL_587319\|20 | ···················· | ···················· | ···················· | ···················· | ···················· | ···················· |
| M | 527 hCoV-19/England/QEUH-A1EBAA/2020\|EPI_ISL_587320\|20 | ···················· | ···················· | ···················· | ···················· | ···················· | ···················· |
| M | 528 hCoV-19/England/QEUH-A1DADB/2020\|EPI_ISL_587321\|20 | ···················· | ···················· | ···················· | ···················· | ···················· | ···················· |
| M | 529 hCoV-19/England/QEUH-A1DC5D/2020\|EPI_ISL_587322\|20 | ···················· | ···················· | ···················· | ···················· | ···················· | ···················· |
| M | 530 hCoV-19/England/QEUH-A1DDE1/2020\|EPI_ISL_587323\|20 | ···················· | ···················· | ···················· | ···················· | ···················· | ···················· |
| M | 531 hCoV-19/England/QEUH-A1DEA2/2020\|EPI_ISL_587324\|20 | ···················· | ···················· | ···················· | ···················· | ···················· | ···················· |
| M | 532 hCoV-19/England/QEUH-A1F013/2020\|EPI_ISL_587325\|20 | ···················· | ·A·················· | ···················· | ···················· | ···················· | ··A················· |
| M | 533 hCoV-19/England/QEUH-A1E973/2020\|EPI_ISL_587326\|20 | ···················· | ···················· | ···················· | ···················· | ···················· | ···················· |
| M | 534 hCoV-19/England/QEUH-A1E4FA/2020\|EPI_ISL_587327\|20 | ···················· | ···················· | ···················· | ···················· | ···················· | ···················· |
| M | 535 hCoV-19/England/QEUH-A1E849/2020\|EPI_ISL_587328\|20 | ···················· | ···················· | ···················· | ···················· | ···················· | ···················· |
| M | 536 hCoV-19/England/QEUH-A1DA80/2020\|EPI_ISL_587329\|20 | ···················· | ···················· | ···················· | ···················· | ···················· | ···················· |
| M | 537 hCoV-19/England/QEUH-A1F4EA/2020\|EPI_ISL_587330\|20 | ···················· | ···················· | ···················· | ···················· | ···················· | ···················· |
| M | 538 hCoV-19/England/QEUH-A21843/2020\|EPI_ISL_587331\|20 | ···················· | ···················· | ···················· | ···················· | ···················· | ···················· |
| M | 539 hCoV-19/England/QEUH-A1E8C1/2020\|EPI_ISL_587332\|20 | ···················· | ···················· | ···················· | ···················· | ···················· | ···················· |
| M | 540 hCoV-19/England/QEUH-A1E9DD/2020\|EPI_ISL_587333\|20 | ···················· | ···················· | ···················· | ···················· | ···················· | ···················· |
| M | 541 hCoV-19/England/QEUH-A1E21E/2020\|EPI_ISL_587334\|20 | ···················· | ···················· | ···················· | ···················· | ···················· | ···················· |
| M | 542 hCoV-19/England/QEUH-A1DCF3/2020\|EPI_ISL_587335\|20 | ···················· | ···················· | ···················· | ···················· | ···················· | ···················· |
| M | 543 hCoV-19/Scotland/QEUH-9F3104/2020\|EPI_ISL_588600\|2 | ···················· | ···················· | ···················· | ···················· | ···················· | ···················· |
| M | 544 hCoV-19/England/QEUH-9F3FDD/2020\|EPI_ISL_588645\|20 | ···················· | ·A·················· | ···················· | ···················· | ···················· | ··A················· |
| M | 545 hCoV-19/England/QEUH-9F2FFC/2020\|EPI_ISL_588646\|20 | ···················· | ······A············· | ···················· | ···················· | ···················· | ···················· |
| M | 546 hCoV-19/England/QEUH-9F2F56/2020\|EPI_ISL_588648\|20 | ···················· | ···················· | ···················· | ···················· | ···················· | ···················· |
| M | 547 hCoV-19/England/QEUH-9F5560/2020\|EPI_ISL_588649\|20 | ···················· | ···················· | ···················· | ···················· | ···················· | ···················· |
| M | 548 hCoV-19/England/QEUH-9F3650/2020\|EPI_ISL_588650\|20 | ···················· | ···················· | ···················· | ···················· | ···················· | ···················· |
| M | 549 hCoV-19/England/QEUH-9F3C5E/2020\|EPI_ISL_588651\|20 | ···················· | ···················· | ···················· | ···················· | ···················· | ···················· |
| M | 550 hCoV-19/England/QEUH-9F4349/2020\|EPI_ISL_588652\|20 | ···················· | ···················· | ···················· | ···················· | ···················· | ···················· |
| M | 551 hCoV-19/England/QEUH-9F3429/2020\|EPI_ISL_588653\|20 | ···················· | ···················· | ···················· | ···················· | ···················· | ···················· |
| M | 552 hCoV-19/England/QEUH-9F377B/2020\|EPI_ISL_588654\|20 | ···················· | ···················· | ···················· | ···················· | ···················· | ···················· |
| M | 553 hCoV-19/England/QEUH-9F417C/2020\|EPI_ISL_588656\|20 | ···················· | ···················· | ···················· | ···················· | ···················· | ···················· |
| M | 554 hCoV-19/England/QEUH-9F340B/2020\|EPI_ISL_588658\|20 | ···················· | ···················· | ···················· | ···················· | ···················· | ···················· |
| M | 555 hCoV-19/England/QEUH-9F425B/2020\|EPI_ISL_588659\|20 | ···················· | ···················· | ···················· | ···················· | ······T············· | ···············A···· |
| M | 556 hCoV-19/England/QEUH-9F4358/2020\|EPI_ISL_588661\|20 | ···················· | ···················· | ···················· | ···················· | ···················· | ···················· |
| M | 557 hCoV-19/England/QEUH-9F3B06/2020\|EPI_ISL_588662\|20 | ···················· | ···················· | ···················· | ···················· | ···················· | ···················· |
| M | 558 hCoV-19/England/QEUH-9F34ED/2020\|EPI_ISL_588664\|20 | ···················· | ···················· | ···················· | ···················· | ···················· | ···················· |
| M | 559 hCoV-19/England/QEUH-9F3E58/2020\|EPI_ISL_588665\|20 | ···················· | ···················· | ···················· | ···················· | ···················· | ···················· |

| | | | | | | | | |
|---|---|---|---|---|---|---|---|---|
| M | 560 | hCoV-19/England/QEUH-9F3614/2020\|EPI_ISL_588666\|20 | ···················· | ···················· | ···················· | ···················· | ···················· | ···················· |
| M | 561 | hCoV-19/England/QEUH-9D0D27/2020\|EPI_ISL_589910\|20 | ···················· | ·A·················· | ···················· | ···················· | ···················· | ···················· |
| M | 562 | hCoV-19/England/QEUH-9D100F/2020\|EPI_ISL_589912\|20 | ···················· | ·A·················· | ···················· | ···················· | ···················· | ···················· |
| M | 563 | hCoV-19/England/QEUH-9D1236/2020\|EPI_ISL_589914\|20 | ···················· | ···················· | ···················· | ···················· | ···················· | ···················· |
| M | 564 | hCoV-19/England/QEUH-9CFB8B/2020\|EPI_ISL_589915\|20 | ···················· | ···················· | ···················· | ···················· | ···················· | ···················· |
| M | 565 | hCoV-19/England/QEUH-9CFF61/2020\|EPI_ISL_589916\|20 | ···················· | ···················· | ···················· | ···················· | ···················· | ···················· |
| M | 566 | hCoV-19/England/QEUH-9CFC97/2020\|EPI_ISL_589918\|20 | ···················· | ·A·················· | ···················· | ···················· | ···················· | ···················· |
| M | 567 | hCoV-19/England/QEUH-9CF954/2020\|EPI_ISL_589919\|20 | ···················· | ···················· | ···················· | ···················· | ···················· | ···················· |
| M | 568 | hCoV-19/England/QEUH-9D00B5/2020\|EPI_ISL_589920\|20 | ···················· | ···················· | ···················· | ···················· | ···················· | ···················· |
| M | 569 | hCoV-19/England/QEUH-9D0AE4/2020\|EPI_ISL_589921\|20 | ···················· | ···················· | ···················· | ···················· | ···················· | ···················· |
| M | 570 | hCoV-19/England/QEUH-9D0510/2020\|EPI_ISL_589922\|20 | ···················· | ···················· | ···················· | ···················· | ···················· | ···················· |
| M | 571 | hCoV-19/England/QEUH-9CFD2B/2020\|EPI_ISL_589923\|20 | ···················· | ···················· | ···················· | ···················· | ···················· | ···················· |
| M | 572 | hCoV-19/England/QEUH-9D0D18/2020\|EPI_ISL_589924\|20 | ···················· | ···················· | ···················· | ···················· | ···················· | ···················· |
| M | 573 | hCoV-19/England/QEUH-9D08EA/2020\|EPI_ISL_589925\|20 | ···················· | ···················· | ···················· | ···················· | ···················· | ···················· |
| M | 574 | hCoV-19/England/QEUH-9CF8DF/2020\|EPI_ISL_589926\|20 | ···················· | ···················· | ···················· | ···················· | ···················· | ···················· |
| M | 575 | hCoV-19/England/QEUH-9D1175/2020\|EPI_ISL_589927\|20 | ···················· | ···················· | ···················· | ···················· | ···················· | ···················· |
| M | 576 | hCoV-19/England/QEUH-9D0F12/2020\|EPI_ISL_589928\|20 | ···················· | ···················· | ···················· | ···················· | ···················· | ···················· |
| M | 577 | hCoV-19/England/QEUH-9CFA51/2020\|EPI_ISL_589929\|20 | ···················· | ···················· | ···················· | ···················· | ···················· | ···················· |
| M | 578 | hCoV-19/England/QEUH-9D0D90/2020\|EPI_ISL_589931\|20 | ···················· | ···················· | ···················· | ···················· | ···················· | ···················· |
| M | 579 | hCoV-19/England/QEUH-9D0176/2020\|EPI_ISL_589976\|20 | ···················· | ···················· | ···················· | ···················· | ·····T·············· | ···················· |
| M | 580 | hCoV-19/England/QEUH-9D0729/2020\|EPI_ISL_589977\|20 | ···················· | ···················· | ···················· | ···················· | ·····T·············· | ···················· |
| M | 581 | hCoV-19/England/QEUH-9CFFBC/2020\|EPI_ISL_589978\|20 | ···················· | ·A······A··········· | ···················· | ···················· | ···················· | ···················· |
| M | 582 | hCoV-19/England/QEUH-9D0A02/2020\|EPI_ISL_589980\|20 | ···················· | ···················· | ···················· | ···················· | ···················· | ···················· |
| M | 583 | hCoV-19/England/QEUH-9D0273/2020\|EPI_ISL_589981\|20 | ···················· | ···················· | ···················· | ···················· | ···················· | ···················· |
| M | 584 | hCoV-19/England/QEUH-9D0291/2020\|EPI_ISL_589982\|20 | ···················· | ···················· | ···················· | ···················· | ···················· | ···················· |
| M | 585 | hCoV-19/England/QEUH-9D0D36/2020\|EPI_ISL_589983\|20 | ···················· | ·A·················· | ···················· | ···················· | ···················· | ··A················· |
| M | 586 | hCoV-19/England/QEUH-9CFF52/2020\|EPI_ISL_589984\|20 | ···················· | ···················· | ···················· | ···················· | ···················· | ···················· |
| M | 587 | hCoV-19/England/QEUH-9D0668/2020\|EPI_ISL_589985\|20 | ···················· | ···················· | ···················· | ···················· | ···················· | ···················· |
| M | 588 | hCoV-19/England/QEUH-9D03AD/2020\|EPI_ISL_589986\|20 | ···················· | ···················· | ···················· | ···················· | ···················· | ···················· |
| M | 589 | hCoV-19/England/QEUH-9CF918/2020\|EPI_ISL_589987\|20 | ···················· | ···················· | ···················· | ···················· | ···················· | ···················· |
| M | 590 | hCoV-19/England/QEUH-9D0A20/2020\|EPI_ISL_589988\|20 | ···················· | ···················· | ···················· | ···················· | ···················· | ···················· |
| M | 591 | hCoV-19/England/QEUH-9D0FC7/2020\|EPI_ISL_589989\|20 | ···················· | ···················· | ···················· | ···················· | ···················· | ···················· |
| M | 592 | hCoV-19/England/QEUH-9CFE82/2020\|EPI_ISL_589990\|20 | ···················· | ···················· | ···················· | ···················· | ···················· | ···················· |
| M | 593 | hCoV-19/England/QEUH-9D0589/2020\|EPI_ISL_589991\|20 | ···················· | ···················· | ···················· | ···················· | ···················· | ···················· |
| M | 594 | hCoV-19/England/QEUH-9CFBF4/2020\|EPI_ISL_589992\|20 | ···················· | ···················· | ···················· | ···················· | ···················· | ···················· |
| M | 595 | hCoV-19/England/QEUH-9CFD1C/2020\|EPI_ISL_589993\|20 | ···················· | ···················· | ···················· | ···················· | ···················· | ···················· |
| M | 596 | hCoV-19/England/QEUH-9D02DD/2020\|EPI_ISL_589995\|20 | ···················· | ·A·················· | ···················· | ···················· | ···················· | ···················· |
| M | 597 | hCoV-19/England/QEUH-9D061D/2020\|EPI_ISL_589996\|20 | ···················· | ···················· | ···················· | ···················· | ···················· | ···················· |
| M | 598 | hCoV-19/England/QEUH-9CFBD6/2020\|EPI_ISL_589997\|20 | ···················· | ···················· | ···················· | ···················· | ···················· | ···················· |
| M | 599 | hCoV-19/Norway/3295/2020\|EPI_ISL_590900\|2020-09-02 | ···················· | ···················· | ···················· | ···················· | ···················· | ···················· |
| M | 600 | hCoV-19/Norway/3278/2020\|EPI_ISL_590901\|2020-09-02 | ···················· | ···················· | ···················· | ···················· | ···················· | ···················· |
| M | 601 | hCoV-19/Norway/3279/2020\|EPI_ISL_590902\|2020-09-02 | ···················· | ···················· | ···················· | ···················· | ···················· | ···················· |
| M | 602 | hCoV-19/Norway/3280/2020\|EPI_ISL_590903\|2020-09-02 | ···················· | ···················· | ···················· | ···················· | ···················· | ···················· |
| M | 603 | hCoV-19/Norway/3262/2020\|EPI_ISL_590904\|2020-08-31 | ···················· | ·A·················· | ···················· | ···················· | ···················· | ···················· |
| M | 604 | hCoV-19/Norway/3263/2020\|EPI_ISL_590905\|2020-08-31 | ···················· | ·A·················· | ···················· | ···················· | ···················· | ···················· |
| M | 605 | hCoV-19/Norway/3264/2020\|EPI_ISL_590906\|2020-08-31 | ···················· | ·A·················· | ···················· | ···················· | ···················· | ···················· |
| M | 606 | hCoV-19/Norway/3267/2020\|EPI_ISL_590907\|2020-09-01 | ···················· | ···················· | ···················· | ···················· | ···················· | ···················· |
| M | 607 | hCoV-19/Norway/3269/2020\|EPI_ISL_590908\|2020-08-23 | ···················· | ···················· | ···················· | ···················· | ···················· | ···················· |
| M | 608 | hCoV-19/Norway/3271/2020\|EPI_ISL_590909\|2020-08-29 | ···················· | ···················· | ···················· | ···················· | ···················· | ···················· |
| M | 609 | hCoV-19/Norway/3272/2020\|EPI_ISL_590910\|2020-08-24 | ···················· | ···················· | ···················· | ···················· | ···················· | ···················· |
| M | 610 | hCoV-19/Norway/3283/2020\|EPI_ISL_590911\|2020-09-07 | ···················· | ···················· | ···················· | ···················· | ···················· | ···················· |
| M | 611 | hCoV-19/Norway/3281/2020\|EPI_ISL_590912\|2020-09-02 | ···················· | ···················· | ···················· | ···················· | ···················· | ···················· |
| M | 612 | hCoV-19/Norway/3275/2020\|EPI_ISL_590913\|2020-08-29 | ···················· | ···················· | ···················· | ···················· | ···················· | ···················· |
| M | 613 | hCoV-19/Norway/2240/2020\|EPI_ISL_590914\|2020-03-27 | ···················· | ···················· | ···················· | ···················· | ···················· | ···················· |
| M | 614 | hCoV-19/Norway/1986/2020\|EPI_ISL_590915\|2020-03-10 | ···················· | ···················· | ···················· | ···················· | ···················· | ···················· |
| M | 615 | hCoV-19/Norway/2086/2020\|EPI_ISL_590916\|2020-03-09 | ···················· | ···················· | ···················· | ···················· | ···················· | ···················· |
| M | 616 | hCoV-19/Norway/2160/2020\|EPI_ISL_590917\|2020-03-17 | ···················· | ···················· | ···················· | ···················· | ···················· | ···················· |
| M | 617 | hCoV-19/Norway/2358/2020\|EPI_ISL_590918\|2020-03-30 | ···················· | ···················· | ···················· | ···················· | ···················· | ···················· |
| M | 618 | hCoV-19/Norway/3248/2020\|EPI_ISL_590919\|2020-08-05 | ···················· | ···················· | ···················· | ···················· | ···················· | ···················· |
| M | 619 | hCoV-19/Norway/4004/2020\|EPI_ISL_590960\|2020-09-21 | ···················· | ···················· | ···················· | ···················· | ···················· | ···················· |
| M | 620 | hCoV-19/Norway/4006/2020\|EPI_ISL_590961\|2020-09-21 | ···················· | ···················· | ···················· | ···················· | ···················· | ···················· |
| M | 621 | hCoV-19/Norway/4007/2020\|EPI_ISL_590962\|2020-09-21 | ···················· | ···················· | ···················· | ···················· | ···················· | ···················· |
| M | 622 | hCoV-19/Norway/4010/2020\|EPI_ISL_590963\|2020-09-21 | ···················· | ···················· | ···················· | ···················· | ···················· | ···················· |
| M | 623 | hCoV-19/Norway/4011/2020\|EPI_ISL_590964\|2020-09-21 | ···················· | ···················· | ···················· | ···················· | ···················· | ···················· |
| M | 624 | hCoV-19/Norway/4012/2020\|EPI_ISL_590965\|2020-09-21 | ···················· | ···················· | ···················· | ···················· | ···················· | ···················· |
| M | 625 | hCoV-19/Norway/4013/2020\|EPI_ISL_590966\|2020-09-21 | ···················· | ···················· | ···················· | ···················· | ···················· | ···················· |
| M | 626 | hCoV-19/Norway/4014/2020\|EPI_ISL_590967\|2020-09-21 | ···················· | ···················· | ···················· | ···················· | ···················· | ···················· |
| M | 627 | hCoV-19/Norway/4015/2020\|EPI_ISL_590968\|2020-09-21 | ···················· | ···················· | ···················· | ···················· | ···················· | ···················· |
| M | 628 | hCoV-19/Norway/4018/2020\|EPI_ISL_590969\|2020-09-21 | ···················· | ···················· | ···················· | ···················· | ···················· | ···················· |
| M | 629 | hCoV-19/Norway/4019/2020\|EPI_ISL_590970\|2020-09-21 | ···················· | ···················· | ···················· | ···················· | ···················· | ···················· |
| M | 630 | hCoV-19/Norway/4021/2020\|EPI_ISL_590971\|2020-09-21 | ···················· | ···················· | ···················· | ···················· | ···················· | ···················· |
| M | 631 | hCoV-19/Norway/4025/2020\|EPI_ISL_590972\|2020-09-21 | ···················· | ···················· | ···················· | ···················· | ···················· | ···················· |
| M | 632 | hCoV-19/Norway/4026/2020\|EPI_ISL_590973\|2020-09-21 | ···················· | ···················· | ···················· | ···················· | ···················· | ···················· |
| M | 633 | hCoV-19/Norway/4027/2020\|EPI_ISL_590974\|2020-09-21 | ···················· | ···················· | ···················· | ···················· | ···················· | ···················· |
| M | 634 | hCoV-19/Norway/4028/2020\|EPI_ISL_590975\|2020-09-21 | ···················· | ···················· | ···················· | ···················· | ···················· | ···················· |
| M | 635 | hCoV-19/Norway/3824/2020\|EPI_ISL_590976\|2020-09-23 | ···················· | ···················· | ···················· | ···················· | ···················· | ···················· |
| M | 636 | hCoV-19/Norway/3826/2020\|EPI_ISL_590977\|2020-09-24 | ···················· | ·········A·········· | ···················· | ···················· | ···················· | ···················· |
| M | 637 | hCoV-19/Norway/3831/2020\|EPI_ISL_590978\|2020-09-24 | ···················· | ···················· | ···················· | ···················· | ···················· | ···················· |
| M | 638 | hCoV-19/Norway/3832/2020\|EPI_ISL_590979\|2020-09-25 | ···················· | ···················· | ···················· | ···················· | ···················· | ···················· |
| M | 640 | hCoV-19/Australia/VIC15029/2020\|EPI_ISL_593112\|202 | ···················· | ···················· | ···················· | ···················· | ···················· | ···················· |

M 641 hCoV-19/Australia/VIC15030/2020|EPI_ISL_593113|202 ···················· ···················· ····················· ···················· ···················· ····················
M 642 hCoV-19/Australia/VIC15031/2020|EPI_ISL_593114|202 ···················· ···················· ····················· ···················· ···················· ····················
M 644 hCoV-19/Australia/VIC15033/2020|EPI_ISL_593116|202 ···················· ···················· ····················· ···················· ···················· ····················
M 645 hCoV-19/Australia/VIC15034/2020|EPI_ISL_593117|202 ···················· ···················· ····················· ···················· ···················· ····················
M 646 hCoV-19/Australia/VIC15036/2020|EPI_ISL_593118|202 ···················· ···················· ····················· ···················· ··N················· ····················
M 647 hCoV-19/Australia/VIC15038/2020|EPI_ISL_593119|202 ···················· ···················· ····················· ···················· ···················· ····················
M 648 hCoV-19/Australia/VIC15043/2020|EPI_ISL_593120|202 ···················· ···················· ····················· ···················· ···················· ····················
M 649 hCoV-19/Australia/VIC15046/2020|EPI_ISL_593121|202 ···················· ···················· ····················· ···················· ···················· ····················
M 650 hCoV-19/Australia/VIC15047/2020|EPI_ISL_593122|202 ···················· ···················· ····················· ···················· ···················· ····················
M 651 hCoV-19/Australia/VIC15049/2020|EPI_ISL_593123|202 ···················· ···················· ····················· ···················· ···················· ····················
M 652 hCoV-19/Australia/VIC15050/2020|EPI_ISL_593124|202 ···················· ···················· ····················· ···················· ···················· ····················
M 653 hCoV-19/Australia/VIC15052/2020|EPI_ISL_593125|202 ···················· ···················· ····················· ···················· ···················· ····················
M 654 hCoV-19/Australia/VIC15069/2020|EPI_ISL_593126|202 ···················· ···················· ····················· ···················· ···················· ····················
M 657 hCoV-19/Australia/VIC15083/2020|EPI_ISL_593129|202 ···················· ···················· ····················· ···················· ···················· ····················
M 658 hCoV-19/Australia/VIC15084/2020|EPI_ISL_593130|202 ···················· ···················· ····················· ···················· ···················· ····················
M 660 hCoV-19/Australia/VIC15088/2020|EPI_ISL_593132|202 ···················· ···················· ····················· ···················· ···················· ····················
M 661 hCoV-19/Australia/VIC15094/2020|EPI_ISL_593133|202 ···················· ···················· ····················· ···················· ···················· ····················
M 662 hCoV-19/Australia/VIC15095/2020|EPI_ISL_593134|202 ···················· ···················· ····················· ···················· ···················· ····················
M 663 hCoV-19/Australia/VIC15096/2020|EPI_ISL_593135|202 ···················· ···················· ····················· ···················· ···················· ····················
M 665 hCoV-19/Australia/VIC15101/2020|EPI_ISL_593137|202 ···················· ···················· ····················· ···················· ···················· ····················
M 667 hCoV-19/Australia/VIC15108/2020|EPI_ISL_593139|202 ···················· ···················· ····················· ···················· ···················· ····················
M 668 hCoV-19/Australia/VIC15110/2020|EPI_ISL_593140|202 ···················· ···················· ····················· ···················· ··········M········· ····················
M 669 hCoV-19/Australia/VIC15111/2020|EPI_ISL_593141|202 ···················· ···················· ····················· ···················· ···················· ····················
M 670 hCoV-19/Australia/VIC15112/2020|EPI_ISL_593142|202 ···················· ···················· ····················· ···················· ···················· ····················
M 671 hCoV-19/Australia/VIC15113/2020|EPI_ISL_593143|202 ···················· ···················· ····················· ···················· ···················· ····················
M 672 hCoV-19/Australia/VIC15114/2020|EPI_ISL_593144|202 ···················· ···················· ····················· ···················· ···················· ····················
M 674 hCoV-19/Australia/VIC15120/2020|EPI_ISL_593146|202 ···················· ···················· ····················· ···················· ···················· ····················
M 675 hCoV-19/Australia/VIC15121/2020|EPI_ISL_593147|202 ···················· ···················· ····················· ···················· ···················· ····················
M 676 hCoV-19/Australia/VIC15122/2020|EPI_ISL_593148|202 ···················· ···················· ····················· ···················· ···················· ····················
M 677 hCoV-19/Australia/VIC15125/2020|EPI_ISL_593149|202 ···················· ···················· ····················· ···················· ···················· ····················
M 678 hCoV-19/Australia/VIC15127/2020|EPI_ISL_593150|202 ···················· ···················· ····················· ···················· ···················· ····················
M 679 hCoV-19/Australia/VIC15129/2020|EPI_ISL_593151|202 ···················· ···················· ····················· ···················· ···················· ····················
M 680 hCoV-19/Australia/VIC15131/2020|EPI_ISL_593152|202 ···················· ···················· ····················· ···················· ···················· ····················
M 681 hCoV-19/Australia/VIC15133/2020|EPI_ISL_593153|202 ···················· ···················· ····················· ···················· ···················· ····················
M 683 hCoV-19/Australia/VIC15154/2020|EPI_ISL_593162|202 ···················· ···················· ····················· ···················· ···················· ····················
M 684 hCoV-19/Australia/VIC15155/2020|EPI_ISL_593163|202 ···················· ···················· ····················· ···················· ···················· ····················
M 686 hCoV-19/Australia/VIC15159/2020|EPI_ISL_593165|202 ···················· ···················· ····················· ···················· ···················· ····················
M 687 hCoV-19/Australia/VIC15186/2020|EPI_ISL_593177|202 ···················· ···················· ····················· ···················· ···················· ····················
M 688 hCoV-19/Australia/VIC15187/2020|EPI_ISL_593178|202 ···················· ···················· ····················· ···················· ···················· ····················
M 689 hCoV-19/Australia/VIC15192/2020|EPI_ISL_593179|202 ···················· ···················· ····················· ···················· ···················· ····················
M 690 hCoV-19/Australia/VIC15193/2020|EPI_ISL_593180|202 ···················· ···················· ····················· ···················· ···················· ····················
M 691 hCoV-19/Australia/VIC15194/2020|EPI_ISL_593181|202 ···················· ···················· ····················· ···················· ···················· ····················
M 692 hCoV-19/Australia/VIC15196/2020|EPI_ISL_593182|202 ···················· ···················· ····················· ···················· ···················· ····················
M 693 hCoV-19/Australia/VIC15197/2020|EPI_ISL_593183|202 ···················· ···················· ····················· ···················· ···················· ····················
M 694 hCoV-19/Australia/VIC15198/2020|EPI_ISL_593184|202 ···················· ···················· ····················· ···················· ···················· ····················
M 695 hCoV-19/Australia/VIC15199/2020|EPI_ISL_593185|202 ···················· ···················· ····················· ···················· ···················· ····················
M 696 hCoV-19/Australia/VIC15202/2020|EPI_ISL_593186|202 ···················· ···················· ····················· ···················· ···················· ····················
M 697 hCoV-19/Australia/VIC15203/2020|EPI_ISL_593187|202 ···················· ···················· ····················· ···················· ···················· ····················
M 698 hCoV-19/Australia/VIC15205/2020|EPI_ISL_593188|202 ···················· ···················· ····················· ···················· ···················· ····················
M 699 hCoV-19/Australia/VIC15206/2020|EPI_ISL_593189|202 ···················· ···················· ····················· ···················· ···················· ····················
M 700 hCoV-19/Australia/VIC15208/2020|EPI_ISL_593190|202 ···················· ···················· ····················· ···················· ···················· ····················
M 701 hCoV-19/Australia/VIC15210/2020|EPI_ISL_593191|202 ···················· ···················· ····················· ···················· ···················· ····················
M 703 hCoV-19/Australia/VIC15214/2020|EPI_ISL_593193|202 ···················· ···················· ····················· ···················· ···················· ····················
M 704 hCoV-19/Australia/VIC15215/2020|EPI_ISL_593194|202 ···················· ···················· ····················· ···················· ···················· ····················
M 706 hCoV-19/Australia/VIC15217/2020|EPI_ISL_593196|202 ···················· ···················· ····················· ···················· ···················· ····················
M 707 hCoV-19/Australia/VIC15218/2020|EPI_ISL_593197|202 ···················· ···················· ····················· ···················· ···················· ····················
M 708 hCoV-19/Australia/VIC15219/2020|EPI_ISL_593198|202 ···················· ···················· ····················· ···················· ···················· ····················
M 709 hCoV-19/USA/UT-UPHL-201014479/2020|EPI_ISL_594442| ···················· ···················· ····················· ···················· ···················· ····················
M 710 hCoV-19/USA/UT-UPHL-201008997/2020|EPI_ISL_594443| ···················· ···················· ····················· ···················· ···················· ····················
M 711 hCoV-19/USA/UT-UPHL-201007233/2020|EPI_ISL_594444| ···················· ···················· ····················· ···················· ···················· ····················
M 712 hCoV-19/USA/WA-CDC-8377/2020|EPI_ISL_594450|2020-0 ···················· ···················· ····················· ···················· ···················· ····················
M 713 hCoV-19/USA/WA-CDC-8378/2020|EPI_ISL_594451|2020-0 ···················· ···················· ····················· ···················· ···················· ····················
M 714 hCoV-19/USA/WA-CDC-8379/2020|EPI_ISL_594452|2020-0 ···················· ···················· ····················· ···················· ···················· ····················
M 715 hCoV-19/USA/WA-CDC-8380/2020|EPI_ISL_594453|2020-0 ···················· ···················· ····················· ···················· ···················· ····················
M 716 hCoV-19/USA/WA-CDC-8381/2020|EPI_ISL_594454|2020-0 ···················· ···················· ····················· ···················· ···················· ····················
M 717 hCoV-19/USA/GA-CDC-8427/2020|EPI_ISL_594456|2020-0 ···················· ···················· ····················· ···················· ···················· ····················
M 718 hCoV-19/USA/NY-CDC-8449/2020|EPI_ISL_594457|2020-0 ···················· ···················· ····················· ···················· ···················· ····················
M 719 hCoV-19/USA/CA-CDC-8452/2020|EPI_ISL_594458|2020-0 ···················· ···················· ····················· ···················· ···················· ····················
M 720 hCoV-19/USA/WA-CDC-8384/2020|EPI_ISL_594459|2020-0 ···················· ···················· ····················· ···················· ···················· ····················
M 721 hCoV-19/USA/DC-CDC-8465/2020|EPI_ISL_594460|2020-0 ···················· ···················· ····················· ···················· ···················· ····················
M 722 hCoV-19/USA/NM-CDC-3728/2020|EPI_ISL_594462|2020-0 ···················· ···················· ····················· ···················· ···················· ····················
M 723 hCoV-19/USA/MN-CDC-3750/2020|EPI_ISL_594463|2020-0 ···················· ···················· ····················· ···················· ···················· ····················
M 724 hCoV-19/USA/FL-CDC-3811/2020|EPI_ISL_594465|2020-0 ···················· ···················· ····················· ···················· ···················· ····················
M 725 hCoV-19/England/ALDP-9E7428/2020|EPI_ISL_594468|20 ···················· ···················· ····················· ···················· ···················· ····················
M 726 hCoV-19/England/ALDP-9E7437/2020|EPI_ISL_594469|20 ···················· ···················· ····················· ···················· ···················· ····················
M 727 hCoV-19/England/ALDP-9E7464/2020|EPI_ISL_594470|20 ···················· ···················· ····················· ···················· ···················· ····················
M 728 hCoV-19/England/ALDP-9E7482/2020|EPI_ISL_594472|20 ···················· ···················· ····················· ···················· ···················· ····················
M 729 hCoV-19/England/ALDP-9E74BF/2020|EPI_ISL_594474|20 ···················· ···················· ····················· ···················· ···················· ····················
M 730 hCoV-19/England/ALDP-9E74CE/2020|EPI_ISL_594475|20 ···················· ···················· ····················· ···················· ···················· ····················
M 731 hCoV-19/England/ALDP-9E7543/2020|EPI_ISL_594477|20 ···················· ···················· ····················· ···················· ···················· ····················

| | | | | | | | | | |
|---|---|---|---|---|---|---|---|---|---|
| M | 732 | hCoV-19/England/ALDP-9E759E/2020\|EPI_ISL_594478\|20 | .................... | .................... | .................... | .................... | .................... | .................... | |
| M | 733 | hCoV-19/England/ALDP-9E75BC/2020\|EPI_ISL_594479\|20 | .................... | .................... | .................... | .................... | .................... | .................... | |
| M | 734 | hCoV-19/England/ALDP-9E75CB/2020\|EPI_ISL_594480\|20 | .................... | .................... | .................... | .................... | ......T............. | .................... | |
| M | 735 | hCoV-19/England/ALDP-9E75DA/2020\|EPI_ISL_594481\|20 | .................... | .................... | .................... | .................... | .................... | .................... | |
| M | 736 | hCoV-19/England/ALDP-9E75E9/2020\|EPI_ISL_594482\|20 | .................... | .................... | .................... | .................... | .................... | .................... | |
| M | 737 | hCoV-19/England/ALDP-9E7604/2020\|EPI_ISL_594483\|20 | .................... | .A.................. | .................... | .................... | .................... | ...A................ | |
| M | 740 | hCoV-19/England/BIRM-64389/2020\|EPI_ISL_594491\|202 | .................... | .A.................. | .................... | .................... | .................... | ...A................ | |
| M | 742 | hCoV-19/England/BIRM-645CF/2020\|EPI_ISL_594496\|202 | .................... | .................... | .................... | .................... | .................... | .................... | |
| M | 743 | hCoV-19/England/QEUH-9F9ECB/2020\|EPI_ISL_595773\|20 | .................... | .................... | .................... | .................... | .................... | .................... | |
| M | 744 | hCoV-19/England/QEUH-9F9EF8/2020\|EPI_ISL_595774\|20 | .................... | .................... | .................... | .................... | .................... | .................... | |
| M | 745 | hCoV-19/Wales/QEUH-9F9F04/2020\|EPI_ISL_595775\|2020 | .................... | .................... | .................... | .................... | .................... | .................... | |
| M | 746 | hCoV-19/England/QEUH-9F9F13/2020\|EPI_ISL_595776\|20 | .................... | .................... | .................... | .................... | .................... | .................... | |
| M | 747 | hCoV-19/England/QEUH-9F9F22/2020\|EPI_ISL_595777\|20 | .................... | .................... | .................... | .................... | ......T............. | .................... | |
| M | 748 | hCoV-19/England/QEUH-9F9F31/2020\|EPI_ISL_595778\|20 | .................... | .................... | .................... | .................... | .................... | .................... | |
| M | 749 | hCoV-19/England/QEUH-9F9F40/2020\|EPI_ISL_595779\|20 | .................... | .................... | .................... | .................... | .................... | .................... | |
| M | 750 | hCoV-19/England/QEUH-9F9F5F/2020\|EPI_ISL_595780\|20 | .................... | .................... | .................... | .................... | .................... | .................... | |
| M | 751 | hCoV-19/England/QEUH-9F9F7D/2020\|EPI_ISL_595781\|20 | .................... | .................... | .................... | .................... | .................... | .................... | |
| M | 752 | hCoV-19/England/QEUH-9F9F9B/2020\|EPI_ISL_595782\|20 | .................... | .................... | .................... | .................... | .................... | .................... | |
| M | 753 | hCoV-19/England/QEUH-9F9FF5/2020\|EPI_ISL_595783\|20 | .................... | .................... | .................... | .................... | .................... | .................... | |
| M | 754 | hCoV-19/England/QEUH-9FA01F/2020\|EPI_ISL_595784\|20 | .................... | .................... | .................... | .................... | .................... | .................... | |
| M | 755 | hCoV-19/England/QEUH-9FA02E/2020\|EPI_ISL_595785\|20 | .................... | .................... | .................... | .................... | .................... | .................... | |
| M | 756 | hCoV-19/England/QEUH-9FA079/2020\|EPI_ISL_595786\|20 | .................... | .................... | .................... | .................... | .................... | .................... | |
| M | 757 | hCoV-19/England/QEUH-9FA097/2020\|EPI_ISL_595787\|20 | .................... | .................... | .................... | .................... | .................... | .................... | |
| M | 758 | hCoV-19/England/QEUH-9FA0A6/2020\|EPI_ISL_595788\|20 | .................... | .................... | .................... | .................... | .................... | .................... | |
| M | 759 | hCoV-19/England/QEUH-9FA13A/2020\|EPI_ISL_595789\|20 | .................... | .................... | .................... | .................... | .................... | .................... | |
| M | 760 | hCoV-19/England/QEUH-9FA167/2020\|EPI_ISL_595790\|20 | .................... | .................... | .................... | .................... | .................... | .................... | |
| M | 761 | hCoV-19/England/QEUH-9FA176/2020\|EPI_ISL_595791\|20 | .................... | .................... | .................... | .................... | .................... | .................... | |
| M | 762 | hCoV-19/England/QEUH-9FA1FE/2020\|EPI_ISL_595792\|20 | .................... | .................... | .................... | .................... | .................... | .................... | |
| M | 763 | hCoV-19/England/QEUH-9FA219/2020\|EPI_ISL_595793\|20 | .................... | .................... | .................... | .................... | .................... | .................... | |
| M | 764 | hCoV-19/England/QEUH-9FA228/2020\|EPI_ISL_595794\|20 | .................... | .................... | .................... | .................... | .................... | .................... | |
| M | 765 | hCoV-19/England/QEUH-9FA255/2020\|EPI_ISL_595795\|20 | .................... | .................... | .................... | .................... | .................... | .................... | |
| M | 766 | hCoV-19/England/QEUH-9FA282/2020\|EPI_ISL_595796\|20 | .................... | .................... | .................... | .................... | .................... | .................... | |
| M | 767 | hCoV-19/England/QEUH-9FA2BF/2020\|EPI_ISL_595797\|20 | .................... | .A.................. | .................... | .................... | .................... | ...A................ | |
| M | 768 | hCoV-19/England/QEUH-9FA2DD/2020\|EPI_ISL_595798\|20 | .................... | .................... | .................... | .................... | .................... | .................... | |
| M | 769 | hCoV-19/Austria/CeMM0585/2020\|EPI_ISL_583601\|2020- | .................... | .................... | .................... | .................... | .................... | .................... | |
| M | 770 | hCoV-19/Austria/CeMM0587/2020\|EPI_ISL_583602\|2020- | .................... | .................... | .................... | .................... | .................... | .................... | |
| M | 771 | hCoV-19/Austria/CeMM0588/2020\|EPI_ISL_583603\|2020- | .................... | .................... | .................... | .................... | .................... | .................... | |
| M | 772 | hCoV-19/Austria/CeMM0590/2020\|EPI_ISL_583605\|2020- | .................... | .................... | .................... | .................... | .................... | .................... | |
| M | 773 | hCoV-19/Austria/CeMM0591/2020\|EPI_ISL_583606\|2020- | .................... | .................... | .................... | .................... | ..N...........NN.... | .................... | 1 |
| M | 774 | hCoV-19/Austria/CeMM0594/2020\|EPI_ISL_583607\|2020- | .................... | .................... | .................... | .................... | .................... | .................... | |
| M | 775 | hCoV-19/Austria/CeMM0595/2020\|EPI_ISL_583608\|2020- | .................... | .................... | .................... | .................... | .................... | .................... | |
| M | 776 | hCoV-19/Austria/CeMM0597/2020\|EPI_ISL_583609\|2020- | .................... | .................... | .................... | .................... | .................... | .................... | |
| M | 777 | hCoV-19/Austria/CeMM0598/2020\|EPI_ISL_583610\|2020- | .................... | .................... | .................... | .................... | .................... | .................... | |
| M | 778 | hCoV-19/Austria/CeMM0600/2020\|EPI_ISL_583611\|2020- | .................... | .................... | .................... | .................... | .................... | .................... | |
| M | 779 | hCoV-19/Austria/CeMM0601/2020\|EPI_ISL_583612\|2020- | .................... | .................... | .................... | .................... | .................... | .................... | |
| M | 780 | hCoV-19/Austria/CeMM0607/2020\|EPI_ISL_583613\|2020- | .................... | .................... | .................... | .................... | .................... | .................... | |
| M | 781 | hCoV-19/Austria/CeMM0608/2020\|EPI_ISL_583614\|2020- | .................... | .................... | .................... | .................... | .................... | .................... | |
| M | 782 | hCoV-19/Austria/CeMM0616/2020\|EPI_ISL_583615\|2020- | .................... | .................... | .................... | .................... | .................... | .................... | |
| M | 783 | hCoV-19/Austria/CeMM0617/2020\|EPI_ISL_583616\|2020- | .................... | .................... | .................... | .................... | .................... | .................... | |
| M | 784 | hCoV-19/Austria/CeMM0642/2020\|EPI_ISL_583619\|2020- | .................... | .................... | .................... | .................... | .................... | .................... | |
| M | 785 | hCoV-19/Austria/CeMM0646/2020\|EPI_ISL_583620\|2020- | .................... | .................... | .................... | .................... | .................... | .................... | |
| M | 786 | hCoV-19/Austria/CeMM0647/2020\|EPI_ISL_583621\|2020- | .................... | .................... | .................... | .................... | .................... | .................... | |
| M | 787 | hCoV-19/Austria/CeMM0648/2020\|EPI_ISL_583622\|2020- | .................... | .................... | .................... | .................... | .................... | .................... | |
| M | 788 | hCoV-19/Austria/CeMM0650/2020\|EPI_ISL_583623\|2020- | .................... | .................... | .................... | .................... | .................... | .................... | |
| M | 789 | hCoV-19/Austria/CeMM0653/2020\|EPI_ISL_583624\|2020- | .................... | .................... | .................... | .................... | .................... | .................... | |
| M | 790 | hCoV-19/Austria/CeMM0654/2020\|EPI_ISL_583625\|2020- | .................... | .................... | .................... | .................... | .................... | .................... | |
| M | 791 | hCoV-19/Austria/CeMM0655/2020\|EPI_ISL_583626\|2020- | .................... | .................... | .................... | .................... | .................... | .................... | |
| M | 792 | hCoV-19/Austria/CeMM0657/2020\|EPI_ISL_583627\|2020- | .................... | .................... | .................... | .................... | .................... | .................... | |
| M | 793 | hCoV-19/Austria/CeMM0660/2020\|EPI_ISL_583629\|2020- | .................... | .................... | .................... | .................... | .................... | .................... | |
| M | 794 | hCoV-19/Austria/CeMM0662/2020\|EPI_ISL_583630\|2020- | .................... | .................... | .................... | .................... | .................... | .................... | |
| M | 795 | hCoV-19/Austria/CeMM0663/2020\|EPI_ISL_583631\|2020- | .................... | .................... | .................... | .................... | .................... | .................... | |
| M | 796 | hCoV-19/Austria/CeMM0667/2020\|EPI_ISL_583633\|2020- | .................... | .................... | .................... | .................... | .................... | .................... | |
| M | 797 | hCoV-19/Austria/CeMM0669/2020\|EPI_ISL_583634\|2020- | .................... | .................... | .................... | .................... | .................... | .................... | |
| M | 798 | hCoV-19/Austria/CeMM0672/2020\|EPI_ISL_583635\|2020- | .................... | .................... | .................... | .................... | .................... | .................... | |
| M | 799 | hCoV-19/Austria/CeMM0673/2020\|EPI_ISL_583636\|2020- | .................... | .................... | .................... | .................... | .................... | .................... | |
| M | 800 | hCoV-19/Austria/CeMM0674/2020\|EPI_ISL_583637\|2020- | .................... | .................... | .................... | .................... | .................... | .................... | |
| M | 801 | hCoV-19/Austria/CeMM0678/2020\|EPI_ISL_583639\|2020- | .................... | .................... | .................... | .................... | .................... | .................... | |
| M | 802 | hCoV-19/Austria/CeMM0679/2020\|EPI_ISL_583640\|2020- | .................... | .................... | .................... | .................... | .................... | .................... | |
| M | 803 | hCoV-19/Austria/CeMM0681/2020\|EPI_ISL_583641\|2020- | .................... | .................... | .................... | .................... | .................... | .................... | |
| M | 804 | hCoV-19/Austria/CeMM0682/2020\|EPI_ISL_583642\|2020- | .................... | .................... | .................... | .................... | .................... | .................... | |
| M | 805 | hCoV-19/Austria/CeMM0683/2020\|EPI_ISL_583643\|2020- | .................... | .................... | .................... | .................... | .................... | .................... | |
| M | 806 | hCoV-19/Austria/CeMM0684/2020\|EPI_ISL_583644\|2020- | .................... | .................... | .................... | .................... | .................... | .................... | |
| M | 807 | hCoV-19/Austria/CeMM0685/2020\|EPI_ISL_583645\|2020- | .................... | .................... | .................... | .................... | .................... | .................... | |
| M | 808 | hCoV-19/Austria/CeMM0686/2020\|EPI_ISL_583646\|2020- | .................... | .................... | .................... | .................... | .................... | .................... | |
| M | 809 | hCoV-19/Austria/CeMM0688/2020\|EPI_ISL_583647\|2020- | .................... | .................... | .................... | .................... | .................... | .................... | |
| M | 810 | hCoV-19/Austria/CeMM0691/2020\|EPI_ISL_583648\|2020- | .................... | .................... | .................... | .................... | .................... | .................... | |
| M | 811 | hCoV-19/Austria/CeMM0692/2020\|EPI_ISL_583649\|2020- | .................... | .................... | .................... | .................... | .................... | .................... | |
| M | 812 | hCoV-19/Austria/CeMM0693/2020\|EPI_ISL_583650\|2020- | .................... | .................... | .................... | .................... | .................... | .................... | |
| M | 813 | hCoV-19/Austria/CeMM0694/2020\|EPI_ISL_583651\|2020- | .................... | .................... | .................... | .................... | .................... | .................... | |
| M | 814 | hCoV-19/Austria/CeMM0696/2020\|EPI_ISL_583652\|2020- | .................... | .................... | .................... | .................... | .................... | .................... | |

| | | | | | | | |
|---|---|---|---|---|---|---|---|
| M | 815 hCoV-19/Austria/CeMM0697/2020\|EPI_ISL_583653\|2020- | ···················· | ···················· | ···················· | ···················· | ··········N········· | ···················· |
| M | 816 hCoV-19/Austria/CeMM0698/2020\|EPI_ISL_583654\|2020- | ···················· | ···················· | ···················· | ···················· | ···················· | ···················· |
| M | 817 hCoV-19/Austria/CeMM0699/2020\|EPI_ISL_583655\|2020- | ···················· | ···················· | ···················· | ···················· | ···················· | ···················· |
| M | 818 hCoV-19/Austria/CeMM0701/2020\|EPI_ISL_583656\|2020- | ···················· | ···················· | ···················· | ···················· | ···················· | ···················· |
| M | 819 hCoV-19/Austria/CeMM0702/2020\|EPI_ISL_583657\|2020- | ···················· | ···················· | ···················· | ···················· | ···················· | ···················· |
| M | 820 hCoV-19/Austria/CeMM0703/2020\|EPI_ISL_583658\|2020- | ···················· | ···················· | ···················· | ···················· | ···················· | ···················· |
| M | 821 hCoV-19/Austria/CeMM0705/2020\|EPI_ISL_583659\|2020- | ···················· | ···················· | ···················· | ···················· | ···················· | ···················· |
| M | 822 hCoV-19/Austria/CeMM0706/2020\|EPI_ISL_583660\|2020- | ···················· | ···················· | ···················· | ···················· | ···················· | ···················· |
| M | 823 hCoV-19/Austria/CeMM0707/2020\|EPI_ISL_583661\|2020- | ···················· | ···················· | ···················· | ···················· | ···················· | ···················· |
| M | 824 hCoV-19/Austria/CeMM0708/2020\|EPI_ISL_583662\|2020- | ···················· | ···················· | ···················· | ···················· | ···················· | ···················· |
| M | 825 hCoV-19/Austria/CeMM0710/2020\|EPI_ISL_583663\|2020- | ···················· | ···················· | ···················· | ···················· | ···················· | ···················· |
| M | 826 hCoV-19/Austria/CeMM0711/2020\|EPI_ISL_583664\|2020- | ···················· | ···················· | ···················· | ···················· | ···················· | ···················· |
| M | 827 hCoV-19/Austria/CeMM0714/2020\|EPI_ISL_583665\|2020- | ···················· | ···················· | ···················· | ···················· | ···················· | ···················· |
| M | 828 hCoV-19/Austria/CeMM0717/2020\|EPI_ISL_583666\|2020- | ···················· | ···················· | ···················· | ···················· | ···················· | ···················· |
| M | 829 hCoV-19/Austria/CeMM0718/2020\|EPI_ISL_583667\|2020- | ···················· | ···················· | ···················· | ···················· | ···················· | ···················· |
| M | 830 hCoV-19/Austria/CeMM0719/2020\|EPI_ISL_583668\|2020- | ···················· | ···················· | ···················· | ···················· | ···················· | ···················· |
| M | 831 hCoV-19/Austria/CeMM0722/2020\|EPI_ISL_583669\|2020- | ···················· | ···················· | ···················· | ···················· | ···················· | ···················· |
| M | 832 hCoV-19/Austria/CeMM0726/2020\|EPI_ISL_583670\|2020- | ···················· | ···················· | ···················· | ···················· | ···················· | ···················· |
| M | 833 hCoV-19/Austria/CeMM0727/2020\|EPI_ISL_583671\|2020- | ···················· | ···················· | ···················· | ···················· | ···················· | ···················· |
| M | 834 hCoV-19/Austria/CeMM0728/2020\|EPI_ISL_583672\|2020- | ···················· | ···················· | ···················· | ···················· | ···················· | ···················· |
| M | 839 hCoV-19/England/ALDP-A2EB5C/2020\|EPI_ISL_587116\|20 | ···················· | ···················· | ···················· | ···················· | ···················· | ···················· |
| M | 840 hCoV-19/England/ALDP-A2F03E/2020\|EPI_ISL_587117\|20 | ···················· | ···················· | ···················· | ···················· | ···················· | ···················· |
| M | 841 hCoV-19/England/ALDP-A2F0F2/2020\|EPI_ISL_587118\|20 | ···················· | ···················· | ···················· | ···················· | ···················· | ···················· |
| M | 842 hCoV-19/England/ALDP-A2F57B/2020\|EPI_ISL_587119\|20 | ···················· | ···················· | ···················· | ···················· | ···················· | ···················· |
| M | 843 hCoV-19/England/ALDP-A2FC85/2020\|EPI_ISL_587120\|20 | ···················· | ···················· | ···················· | ···················· | ···················· | ···················· |
| M | 844 hCoV-19/England/ALDP-A2E758/2020\|EPI_ISL_587121\|20 | ···················· | ···················· | ···················· | ···················· | ···················· | ···················· |
| M | 845 hCoV-19/England/ALDP-A2EF05/2020\|EPI_ISL_587122\|20 | ···················· | ···················· | ···················· | ···················· | ···················· | ···················· |
| M | 846 hCoV-19/England/ALDP-A2EFBA/2020\|EPI_ISL_587123\|20 | ···················· | ···················· | ···················· | ···················· | ···················· | ···················· |
| M | 847 hCoV-19/England/ALDP-A2F62D/2020\|EPI_ISL_587125\|20 | ···················· | ···················· | ···················· | ···················· | ···················· | ···················· |
| M | 848 hCoV-19/England/ALDP-A2F6D2/2020\|EPI_ISL_587126\|20 | ···················· | ···················· | ···················· | ···················· | ···················· | ···················· |
| M | 849 hCoV-19/England/ALDP-A2F9F7/2020\|EPI_ISL_587138\|20 | ···················· | ···················· | ···················· | ···················· | ···················· | ···················· |
| M | 850 hCoV-19/England/ALDP-A2E864/2020\|EPI_ISL_587139\|20 | ···················· | ·A·················· | ···················· | ···················· | ···················· | ···A················ |
| M | 851 hCoV-19/England/ALDP-A2F001/2020\|EPI_ISL_587140\|20 | ···················· | ···················· | ···················· | ···················· | ···················· | ···················· |
| M | 852 hCoV-19/England/ALDP-A2FB79/2020\|EPI_ISL_587141\|20 | ···················· | ···················· | ···················· | ···················· | ···················· | ···················· |
| M | 853 hCoV-19/England/ALDP-A2F72A/2020\|EPI_ISL_587142\|20 | ···················· | ···················· | ···················· | ···················· | ···················· | ···················· |
| M | 854 hCoV-19/England/ALDP-A2EC2C/2020\|EPI_ISL_587143\|20 | ···················· | ·A·················· | ···················· | ···················· | ···················· | ···A················ |
| M | 855 hCoV-19/England/ALDP-A2F6A5/2020\|EPI_ISL_587144\|20 | ···················· | ···················· | ···················· | ···················· | ···················· | ···················· |
| M | 856 hCoV-19/England/ALDP-A2FCB2/2020\|EPI_ISL_587145\|20 | ···················· | ···················· | ···················· | ···················· | ···················· | ···················· |
| M | 857 hCoV-19/England/ALDP-A2F14A/2020\|EPI_ISL_587146\|20 | ···················· | ·A·················· | ···················· | ···················· | ···················· | ···A················ |
| M | 858 hCoV-19/England/ALDP-A2EE08/2020\|EPI_ISL_587148\|20 | ···················· | ···················· | ···················· | ···················· | ···················· | ···················· |
| M | 859 hCoV-19/England/ALDP-A2F775/2020\|EPI_ISL_587149\|20 | ···················· | ···················· | ···················· | ···················· | ···················· | ···················· |
| M | 860 hCoV-19/England/ALDP-A2EE17/2020\|EPI_ISL_587150\|20 | ···················· | ···················· | ···················· | ···················· | ···················· | ···················· |
| M | 861 hCoV-19/Wales/ALDP-A2FAD6/2020\|EPI_ISL_587151\|2020 | ···················· | ···················· | ···················· | ···················· | ···················· | ···················· |
| M | 862 hCoV-19/England/ALDP-A2F511/2020\|EPI_ISL_587152\|20 | ···················· | ···················· | ···················· | ···················· | ···················· | ···················· |
| M | 863 hCoV-19/England/ALDP-A2F11D/2020\|EPI_ISL_587153\|20 | ···················· | ···················· | ···················· | ···················· | ···················· | ···················· |
| M | 864 hCoV-19/England/ALDP-A2FBA6/2020\|EPI_ISL_587154\|20 | ···················· | ···················· | ···················· | ···················· | ···················· | ···················· |
| M | 865 hCoV-19/England/ALDP-A2F748/2020\|EPI_ISL_587155\|20 | ···················· | ···················· | ···················· | ···················· | ······T············· | ···················· |
| M | 866 hCoV-19/England/ALDP-A2EE44/2020\|EPI_ISL_587156\|20 | ···················· | ···················· | ···················· | ···················· | ···················· | ···················· |
| M | 867 hCoV-19/England/ALDP-A2F836/2020\|EPI_ISL_587157\|20 | ···················· | ···················· | ···················· | ···················· | ···················· | ···················· |
| M | 868 hCoV-19/England/ALDP-A2EA31/2020\|EPI_ISL_587158\|20 | ···················· | ···················· | ···················· | ···················· | ···················· | ···················· |
| M | 869 hCoV-19/England/ALDP-A2F159/2020\|EPI_ISL_587159\|20 | ···················· | ···················· | ···················· | ···················· | ···················· | ···················· |
| M | 870 hCoV-19/England/QEUH-9F4B32/2020\|EPI_ISL_588447\|20 | ···················· | ···················· | ···················· | ···················· | ···················· | ···················· |
| M | 871 hCoV-19/England/QEUH-9F5937/2020\|EPI_ISL_588448\|20 | ···················· | ···················· | ···················· | ···················· | ···················· | ···················· |
| M | 872 hCoV-19/England/QEUH-9F58EF/2020\|EPI_ISL_588449\|20 | ···················· | ···················· | ···················· | ···················· | ···················· | ···················· |
| M | 873 hCoV-19/England/QEUH-9F481D/2020\|EPI_ISL_588450\|20 | ···················· | ···················· | ···················· | ···················· | ···················· | ···················· |
| M | 874 hCoV-19/England/QEUH-9F58B2/2020\|EPI_ISL_588451\|20 | ···················· | ···················· | ···················· | ···················· | ···················· | ···················· |
| M | 875 hCoV-19/England/QEUH-9F476B/2020\|EPI_ISL_588452\|20 | ···················· | ·A·················· | ···················· | ···················· | ···················· | ···················· |
| M | 876 hCoV-19/England/QEUH-9F5E38/2020\|EPI_ISL_588453\|20 | ···················· | ···················· | ···················· | ···················· | ···················· | ···················· |
| M | 877 hCoV-19/England/QEUH-9F5D1D/2020\|EPI_ISL_588454\|20 | ···················· | ···················· | ···················· | ···················· | ···················· | ···················· |
| M | 878 hCoV-19/England/QEUH-9F5005/2020\|EPI_ISL_588456\|20 | ···················· | ···················· | ···················· | ···················· | ···················· | ···················· |
| M | 879 hCoV-19/England/QEUH-9F6611/2020\|EPI_ISL_588457\|20 | ···················· | ···················· | ···················· | ···················· | ···················· | ···················· |
| M | 880 hCoV-19/England/QEUH-9F4ADB/2020\|EPI_ISL_588469\|20 | ···················· | ···················· | ···················· | ···················· | ···················· | ···················· |
| M | 881 hCoV-19/England/QEUH-9F474D/2020\|EPI_ISL_588472\|20 | ···················· | ···················· | ···················· | ···················· | ···················· | ···················· |
| M | 882 hCoV-19/England/QEUH-9F4E66/2020\|EPI_ISL_588473\|20 | ···················· | ···················· | ···················· | ···················· | ···················· | ···················· |
| M | 883 hCoV-19/England/QEUH-9F51F3/2020\|EPI_ISL_588474\|20 | ···················· | ···················· | ···················· | ···················· | ···················· | ···················· |
| M | 884 hCoV-19/England/QEUH-9F524B/2020\|EPI_ISL_588475\|20 | ···················· | ···················· | ···················· | ···················· | ···················· | ···················· |
| M | 885 hCoV-19/England/QEUH-9F416D/2020\|EPI_ISL_588476\|20 | ···················· | ···················· | ···················· | ···················· | ···················· | ···················· |
| M | 886 hCoV-19/England/QEUH-9F34DE/2020\|EPI_ISL_588477\|20 | ···················· | ···················· | ···················· | ···················· | ······T············· | ···················· |
| M | 887 hCoV-19/England/QEUH-9F33C2/2020\|EPI_ISL_588478\|20 | ···················· | ···················· | ···················· | ···················· | ···················· | ···················· |
| M | 888 hCoV-19/England/QEUH-9F37A8/2020\|EPI_ISL_588479\|20 | ···················· | ···················· | ···················· | ···················· | ···················· | ···················· |
| M | 889 hCoV-19/England/QEUH-9F35BD/2020\|EPI_ISL_588481\|20 | ···················· | ···················· | ···················· | ···················· | ···················· | ···················· |
| M | 890 hCoV-19/England/QEUH-9F317D/2020\|EPI_ISL_588482\|20 | ···················· | ···················· | ···················· | ···················· | ···················· | ···················· |
| M | 891 hCoV-19/England/QEUH-9F34B0/2020\|EPI_ISL_588483\|20 | ···················· | ···················· | ···················· | ···················· | ···················· | ···················· |
| M | 892 hCoV-19/Northern Ireland/QEUH-9F32D4/2020\|EPI_ISL_ | ···················· | ···················· | ···················· | ···················· | ···················· | ···················· |
| M | 893 hCoV-19/England/QEUH-9F2E1D/2020\|EPI_ISL_588485\|20 | ···················· | ···················· | ···················· | ···················· | ····T··············· | ···················· |
| M | 894 hCoV-19/England/QEUH-9F3AEB/2020\|EPI_ISL_588486\|20 | ···················· | ···················· | ···················· | ···················· | ···················· | ···················· |
| M | 895 hCoV-19/England/QEUH-9F3359/2020\|EPI_ISL_588487\|20 | ···················· | ···················· | ···················· | ···················· | ···················· | ···················· |
| M | 896 hCoV-19/England/QEUH-9F4112/2020\|EPI_ISL_588488\|20 | ···················· | ···················· | ···················· | ···················· | ···················· | ···················· |
| M | 897 hCoV-19/England/QEUH-9F330E/2020\|EPI_ISL_588489\|20 | ···················· | ···················· | ···················· | ···················· | ···················· | ···················· |
| M | 898 hCoV-19/England/ALDP-9EA4F8/2020\|EPI_ISL_589778\|20 | ···················· | ···················· | ···················· | ···················· | ···················· | ···················· |

| | | | | | | | | |
|---|---|---|---|---|---|---|---|---|
| M | 899 | hCoV-19/England/ALDP-9EAB99/2020\|EPI_ISL_589779\|20 | ···················· | ···················· | ····················· | ···················· | ···················· | ···················· |
| M | 900 | hCoV-19/Wales/ALDP-9EA0D6/2020\|EPI_ISL_589781\|2020 | ···················· | ···················· | ····················· | ···················· | ···················· | ···················· |
| M | 901 | hCoV-19/England/ALDP-9EADC0/2020\|EPI_ISL_589783\|20 | ···················· | ···················· | ····················· | ···················· | ···················· | ···················· |
| M | 902 | hCoV-19/Wales/ALDP-9EB002/2020\|EPI_ISL_589784\|2020 | ···················· | ···················· | ····················· | ···················· | ···················· | ···················· |
| M | 903 | hCoV-19/Wales/ALDP-9EA09A/2020\|EPI_ISL_589785\|2020 | ···················· | ···················· | ····················· | ···················· | ····T··············· | ···················· |
| M | 904 | hCoV-19/England/ALDP-9EAE45/2020\|EPI_ISL_589786\|20 | ···················· | ···················· | ····················· | ···················· | ···················· | ···················· |
| M | 905 | hCoV-19/Wales/MILK-9E1109/2020\|EPI_ISL_589787\|2020 | ···················· | ·A·················· | ····················· | ···················· | ···················· | ··A················· |
| M | 906 | hCoV-19/England/QEUH-9DC778/2020\|EPI_ISL_589788\|20 | ···················· | ···················· | ····················· | ···················· | ···················· | ···················· |
| M | 907 | hCoV-19/England/QEUH-9D070B/2020\|EPI_ISL_589900\|20 | ···················· | ···················· | ····················· | ···················· | ···················· | ···················· |
| M | 908 | hCoV-19/England/QEUH-9D01C1/2020\|EPI_ISL_589901\|20 | ···················· | ···················· | ····················· | ···················· | ···················· | ···················· |
| M | 909 | hCoV-19/England/QEUH-9D0EF7/2020\|EPI_ISL_589902\|20 | ···················· | ···················· | ····················· | ···················· | ···················· | ···················· |
| M | 910 | hCoV-19/England/QEUH-9D05D4/2020\|EPI_ISL_589903\|20 | ···················· | ···················· | ····················· | ···················· | ···················· | ···················· |
| M | 911 | hCoV-19/England/QEUH-9D0F7C/2020\|EPI_ISL_589905\|20 | ···················· | ···················· | ····················· | ···················· | ···················· | ···················· |
| M | 912 | hCoV-19/England/QEUH-9D013A/2020\|EPI_ISL_589907\|20 | ···················· | ···················· | ····················· | ···················· | ····T··············· | ···················· |
| M | 913 | hCoV-19/England/QEUH-9D05C5/2020\|EPI_ISL_589908\|20 | ···················· | ···················· | ····················· | ···················· | ···················· | ···················· |
| M | 914 | hCoV-19/England/QEUH-9D07DE/2020\|EPI_ISL_589909\|20 | ···················· | ·A·················· | ····················· | ···················· | ···················· | ···················· |
| M | 915 | hCoV-19/USA/MN-MDH-1756/2020\|EPI_ISL_590761\|2020-0 | ···················· | ···················· | ····················· | ···················· | ···················· | ···················· |
| M | 916 | hCoV-19/USA/MN-MDH-1757/2020\|EPI_ISL_590762\|2020-0 | ···················· | ···················· | ····················· | ···················· | ···················· | ···················· |
| M | 917 | hCoV-19/USA/MN-MDH-1758/2020\|EPI_ISL_590763\|2020-0 | ···················· | ···················· | ····················· | ···················· | ···················· | ···················· |
| M | 918 | hCoV-19/USA/MN-MDH-1759/2020\|EPI_ISL_590764\|2020-0 | ···················· | ···················· | ····················· | ···················· | ···················· | ···················· |
| M | 919 | hCoV-19/USA/MN-MDH-1778/2020\|EPI_ISL_590780\|2020-0 | ···················· | ···················· | ····················· | ···················· | ···················· | ···················· |
| M | 920 | hCoV-19/USA/MN-MDH-1779/2020\|EPI_ISL_590781\|2020-0 | ···················· | ···················· | ····················· | ···················· | ···················· | ···················· |
| M | 921 | hCoV-19/USA/MN-MDH-1780/2020\|EPI_ISL_590782\|2020-0 | ···················· | ···················· | ····················· | ···················· | ···················· | ···················· |
| M | 922 | hCoV-19/USA/MN-MDH-1781/2020\|EPI_ISL_590783\|2020-0 | ···················· | ···················· | ····················· | ···················· | ···················· | ···················· |
| M | 923 | hCoV-19/USA/MN-MDH-1782/2020\|EPI_ISL_590784\|2020-0 | ···················· | ···················· | ····················· | ···················· | ···················· | ···················· |
| M | 924 | hCoV-19/USA/MN-MDH-1783/2020\|EPI_ISL_590785\|2020-0 | ···················· | ···················· | ····················· | ···················· | ···················· | ···················· |
| M | 925 | hCoV-19/USA/MN-MDH-1784/2020\|EPI_ISL_590786\|2020-0 | ···················· | ···G················ | ····················· | ···················· | ···················· | ···················· |
| M | 926 | hCoV-19/USA/MN-MDH-1785/2020\|EPI_ISL_590787\|2020-0 | ···················· | ···G················ | ····················· | ···················· | ···················· | ···················· |
| M | 927 | hCoV-19/USA/MN-MDH-1786/2020\|EPI_ISL_590788\|2020-0 | ···················· | ···················· | ····················· | ···················· | ···················· | ···················· |
| M | 928 | hCoV-19/USA/MN-MDH-1787/2020\|EPI_ISL_590789\|2020-0 | ···················· | ···················· | ····················· | ···················· | ···················· | ···················· |
| M | 929 | hCoV-19/USA/MN-MDH-1788/2020\|EPI_ISL_590790\|2020-0 | ···················· | ···················· | ····················· | ···················· | ···················· | ···················· |
| M | 930 | hCoV-19/USA/MN-MDH-1789/2020\|EPI_ISL_590791\|2020-0 | ···················· | ···················· | ····················· | ···················· | ·····N·············· | ···················· |
| M | 931 | hCoV-19/USA/MN-MDH-1790/2020\|EPI_ISL_590792\|2020-0 | ···················· | ···················· | ····················· | ···················· | ···················· | ···················· |
| M | 932 | hCoV-19/USA/MN-MDH-1791/2020\|EPI_ISL_590793\|2020-0 | ···················· | ···················· | ····················· | ···················· | ···················· | ···················· |
| M | 933 | hCoV-19/USA/MN-MDH-1792/2020\|EPI_ISL_590794\|2020-0 | ···················· | ···G················ | ····················· | ···················· | ···················· | ···················· |
| M | 934 | hCoV-19/USA/MN-MDH-1793/2020\|EPI_ISL_590795\|2020-0 | ···················· | ···················· | ····················· | ···················· | ···················· | ···················· |
| M | 935 | hCoV-19/USA/MN-MDH-1794/2020\|EPI_ISL_590796\|2020-0 | ···················· | ···················· | ····················· | ···················· | ···················· | ···················· |
| M | 936 | hCoV-19/USA/MN-MDH-1795/2020\|EPI_ISL_590797\|2020-0 | ···················· | ···················· | ····················· | ···················· | ···················· | ···················· |
| M | 937 | hCoV-19/USA/MN-MDH-1796/2020\|EPI_ISL_590798\|2020-0 | ···················· | ···················· | ····················· | ···················· | ···················· | ···················· |
| M | 938 | hCoV-19/USA/MN-MDH-1797/2020\|EPI_ISL_590799\|2020-0 | ···················· | ···················· | ····················· | ···················· | ···················· | ···················· |
| M | 940 | hCoV-19/Australia/VIC15144/2020\|EPI_ISL_593156\|202 | ···················· | ···················· | ····················· | ···················· | ···················· | ···················· |
| M | 941 | hCoV-19/Australia/VIC15145/2020\|EPI_ISL_593157\|202 | ···················· | ···················· | ····················· | ···················· | ···················· | ···················· |
| M | 942 | hCoV-19/Australia/VIC15148/2020\|EPI_ISL_593158\|202 | ···················· | ···················· | ····················· | ···················· | ···················· | ···················· |
| M | 943 | hCoV-19/Australia/VIC15149/2020\|EPI_ISL_593159\|202 | ···················· | ···················· | ····················· | ···················· | ···················· | ···················· |
| M | 944 | hCoV-19/Australia/VIC15150/2020\|EPI_ISL_593160\|202 | ···················· | ···················· | ····················· | ···················· | ···················· | ···················· |
| M | 945 | hCoV-19/Australia/VIC15152/2020\|EPI_ISL_593161\|202 | ···················· | ···················· | ····················· | ···················· | ···················· | ···················· |
| M | 946 | hCoV-19/Australia/VIC15163/2020\|EPI_ISL_593166\|202 | ···················· | ···················· | ····················· | ···················· | ···················· | ···················· |
| M | 948 | hCoV-19/Australia/VIC15165/2020\|EPI_ISL_593168\|202 | ···················· | ···················· | ····················· | ···················· | ···················· | ···················· |
| M | 949 | hCoV-19/Australia/VIC15168/2020\|EPI_ISL_593169\|202 | ···················· | ···················· | ····················· | ···················· | ···················· | ···················· |
| M | 951 | hCoV-19/Australia/VIC15174/2020\|EPI_ISL_593171\|202 | ···················· | ···················· | ····················· | ···················· | ···················· | ···················· |
| M | 952 | hCoV-19/Australia/VIC15176/2020\|EPI_ISL_593172\|202 | ···················· | ···················· | ····················· | ···················· | ···················· | ···················· |
| M | 953 | hCoV-19/Australia/VIC15178/2020\|EPI_ISL_593173\|202 | ···················· | ···················· | ····················· | ···················· | ···················· | ···················· |
| M | 954 | hCoV-19/Australia/VIC15179/2020\|EPI_ISL_593174\|202 | ···················· | ···················· | ····················· | ···················· | ···················· | ···················· |
| M | 955 | hCoV-19/Australia/VIC15183/2020\|EPI_ISL_593175\|202 | ···················· | ···················· | ····················· | ···················· | ···················· | ···················· |
| M | 956 | hCoV-19/Australia/VIC15185/2020\|EPI_ISL_593176\|202 | ···················· | ···················· | ····················· | ···················· | ···················· | ···················· |
| M | 957 | hCoV-19/USA/FL-BPHL-1610/2020\|EPI_ISL_594300\|2020- | ···················· | ···················· | ····················· | ···················· | ···················· | ···················· |
| M | 958 | hCoV-19/USA/FL-BPHL-1611/2020\|EPI_ISL_594301\|2020- | ···················· | ···················· | ····················· | ···················· | ···················· | ···················· |
| M | 959 | hCoV-19/USA/FL-BPHL-1612/2020\|EPI_ISL_594302\|2020- | ···················· | ···················· | ····················· | ···················· | ···················· | ···················· |
| M | 960 | hCoV-19/USA/FL-BPHL-1613/2020\|EPI_ISL_594303\|2020- | ···················· | ···················· | ····················· | ···················· | ···················· | ···················· |
| M | 961 | hCoV-19/USA/FL-BPHL-1614/2020\|EPI_ISL_594304\|2020- | ···················· | ···················· | ····················· | ···················· | ···················· | ···················· |
| M | 962 | hCoV-19/USA/FL-BPHL-1615/2020\|EPI_ISL_594305\|2020- | ···················· | ···················· | ····················· | ···················· | ···················· | ···················· |
| M | 963 | hCoV-19/USA/FL-BPHL-1616/2020\|EPI_ISL_594306\|2020- | ···················· | ···················· | ····················· | ···················· | ···················· | ···················· |
| M | 964 | hCoV-19/USA/FL-BPHL-1617/2020\|EPI_ISL_594307\|2020- | ···················· | ···················· | ····················· | ···················· | ···················· | ···················· |
| M | 965 | hCoV-19/USA/FL-BPHL-1618/2020\|EPI_ISL_594308\|2020- | ···················· | ···················· | ····················· | ···················· | ···················· | ···················· |
| M | 966 | hCoV-19/USA/FL-BPHL-1620/2020\|EPI_ISL_594310\|2020- | ···················· | ···················· | ····················· | ···················· | ···················· | ···················· |
| M | 967 | hCoV-19/USA/FL-BPHL-1621/2020\|EPI_ISL_594311\|2020- | ···················· | ···················· | ····················· | ···················· | ···················· | ···················· |
| M | 968 | hCoV-19/USA/FL-BPHL-1622/2020\|EPI_ISL_594312\|2020- | ···················· | ···················· | ····················· | ···················· | ···················· | ···················· |
| M | 970 | hCoV-19/USA/FL-BPHL-1624/2020\|EPI_ISL_594314\|2020- | ···················· | ···················· | ····················· | ···················· | ···················· | ···················· |
| M | 971 | hCoV-19/USA/FL-BPHL-1626/2020\|EPI_ISL_594316\|2020- | ···················· | ···················· | ····················· | ···················· | ···················· | ···················· |
| M | 973 | hCoV-19/England/BIRM-6435C/2020\|EPI_ISL_594488\|202 | ···················· | ·A·················· | ····················· | ···················· | ···················· | ··A················· |
| M | 977 | hCoV-19/England/BIRM-64608/2020\|EPI_ISL_594499\|202 | ···················· | ···················· | ····················· | ···················· | ···················· | ···················· |
| M | 978 | hCoV-19/Wales/PHWC-47EE69/2020\|EPI_ISL_595601\|2020 | ···················· | ···················· | ····················· | ···················· | ···················· | ···················· |
| M | 979 | hCoV-19/Wales/PHWC-47EE87/2020\|EPI_ISL_595603\|2020 | ···················· | ···················· | ····················· | ···················· | ···················· | ···················· |
| M | 980 | hCoV-19/Wales/PHWC-47EE96/2020\|EPI_ISL_595604\|2020 | ···················· | ···················· | ····················· | ···················· | ···················· | ···················· |
| M | 981 | hCoV-19/Wales/PHWC-47EEA5/2020\|EPI_ISL_595605\|2020 | ···················· | ···················· | ····················· | ···················· | ···················· | ···················· |
| M | 982 | hCoV-19/Wales/PHWC-47EEC3/2020\|EPI_ISL_595607\|2020 | ···················· | ···················· | ····················· | ···················· | ···················· | ···················· |
| M | 983 | hCoV-19/Wales/PHWC-47EEE1/2020\|EPI_ISL_595609\|2020 | ···················· | ···················· | ····················· | ···················· | ···················· | ···················· |
| M | 984 | hCoV-19/Wales/PHWC-47EEF0/2020\|EPI_ISL_595610\|2020 | ···················· | ···················· | ····················· | ···················· | ···················· | ···················· |
| M | 985 | hCoV-19/Wales/PHWC-47EF1B/2020\|EPI_ISL_595612\|2020 | ···················· | ···················· | ····················· | ···················· | ···················· | ···················· |
| M | 986 | hCoV-19/Wales/PHWC-47EF2A/2020\|EPI_ISL_595613\|2020 | ········Y··········· | ···················· | ····················· | ···················· | ···················· | ···················· |

M 987 hCoV-19/Wales/PHWC-47EF39/2020|EPI_ISL_595614|2020 ···················· ···················· ···················· ···················· ···················· ····················
M 989 hCoV-19/Wales/PHWC-47EF57/2020|EPI_ISL_595616|2020 ···················· ···················· ···················· ···················· ···················· ····················
M 990 hCoV-19/England/QEUH-9F6B9A/2020|EPI_ISL_595617|20 ···················· ···················· ···················· ···················· ···················· ····················
M 991 hCoV-19/Scotland/QEUH-9F6BB8/2020|EPI_ISL_595618|2 ···················· ···················· ···················· ···················· ···················· ····················
M 992 hCoV-19/Scotland/QEUH-9F6BE5/2020|EPI_ISL_595619|2 ···················· ···················· ···················· ···················· ···················· ····················
M 993 hCoV-19/Scotland/QEUH-9F6C2E/2020|EPI_ISL_595620|2 ···················· ···················· ···················· ···················· ···················· ····················
M 994 hCoV-19/England/QEUH-9F6C4C/2020|EPI_ISL_595621|20 ···················· ···················· ···················· ···················· ···················· ····················
M 995 hCoV-19/England/QEUH-9F6C97/2020|EPI_ISL_595622|20 ···················· ···················· ···················· ···················· ···················· ····················
M 996 hCoV-19/Scotland/QEUH-9F6CB5/2020|EPI_ISL_595623|2 ···················· ···················· ···················· ···················· ···················· ····················
M 997 hCoV-19/Scotland/QEUH-9F6CD3/2020|EPI_ISL_595624|2 ···················· ···················· ···················· ···················· ···················· ····················
M 998 hCoV-19/Scotland/QEUH-9F6CE2/2020|EPI_ISL_595625|2 ···················· ···················· ···················· ···················· ···················· ····················
M 999 hCoV-19/England/QEUH-9F6D2B/2020|EPI_ISL_595626|20 ···················· ···················· ···················· ···················· ···················· ····················
M 1000 hCoV-19/Scotland/QEUH-9F6D49/2020|EPI_ISL_595627|2 ···················· ···················· ···················· ···················· ···················· ····················
M 1001 hCoV-19/England/QEUH-9F6D58/2020|EPI_ISL_595628|20 ···················· ···················· ···················· ···················· ···················· ····················
M 1002 hCoV-19/England/QEUH-9F6D67/2020|EPI_ISL_595629|20 ···················· ···················· ···················· ···················· ···················· ····················
M 1003 hCoV-19/England/QEUH-9F6D76/2020|EPI_ISL_595630|20 ···················· ···················· ···················· ···················· ···················· ····················
M 1004 hCoV-19/England/QEUH-9F6DB2/2020|EPI_ISL_595631|20 ···················· ···················· ···················· ···················· ···················· ····················
M 1005 hCoV-19/England/QEUH-9F6DC1/2020|EPI_ISL_595632|20 ···················· ···················· ···················· ···················· ···················· ····················
M 1006 hCoV-19/England/QEUH-9F6DD0/2020|EPI_ISL_595633|20 ···················· ···················· ···················· ···················· ···················· ····················
M 1007 hCoV-19/England/QEUH-9F6DEF/2020|EPI_ISL_595634|20 ···················· ···················· ···················· ···················· ···················· ····················
M 1008 hCoV-19/England/QEUH-9F6E19/2020|EPI_ISL_595635|20 ···················· ···················· ···················· ···················· ···················· ····················
M 1009 hCoV-19/England/QEUH-9F6E28/2020|EPI_ISL_595636|20 ···················· ···················· ···················· ···················· ···················· ····················
M 1010 hCoV-19/England/QEUH-9F6E46/2020|EPI_ISL_595637|20 ···················· ···················· ···················· ···················· ···················· ····················
M 1011 hCoV-19/England/QEUH-9F6E73/2020|EPI_ISL_595638|20 ···················· ···················· ···················· ···················· ···················· ····················
M 1012 hCoV-19/England/QEUH-9F6E82/2020|EPI_ISL_595639|20 ···················· ···················· ···················· ···················· ···················· ····················
M 1013 hCoV-19/England/QEUH-9F6E91/2020|EPI_ISL_595640|20 ···················· ···················· ···················· ···················· ···················· ····················
M 1014 hCoV-19/England/QEUH-9F6EA0/2020|EPI_ISL_595641|20 ···················· ···················· ···················· ···················· ···················· ····················
M 1015 hCoV-19/England/QEUH-9F6EBF/2020|EPI_ISL_595642|20 ···················· ···················· ···················· ···················· ···················· ····················
M 1016 hCoV-19/England/QEUH-9F6ECE/2020|EPI_ISL_595643|20 ···················· ···················· ···················· ···················· ···················· ····················
M 1017 hCoV-19/England/QEUH-9F6EFB/2020|EPI_ISL_595644|20 ···················· ···················· ···················· ···················· ···················· ····················
M 1018 hCoV-19/England/QEUH-9F6F07/2020|EPI_ISL_595645|20 ···················· ···················· ···················· ···················· ···················· ····················
M 1019 hCoV-19/England/QEUH-9F6F25/2020|EPI_ISL_595646|20 ···················· ···················· ···················· ···················· ···················· ····················
M 1020 hCoV-19/England/QEUH-9F6F43/2020|EPI_ISL_595647|20 ···················· ···················· ···················· ···················· ···················· ····················
M 1021 hCoV-19/England/QEUH-9F6F8F/2020|EPI_ISL_595648|20 ···················· ···················· ···················· ···················· ···················· ····················
M 1022 hCoV-19/Scotland/QEUH-9F6FAD/2020|EPI_ISL_595649|2 ···················· ···················· ···················· ···················· ···················· ····················
M 1023 hCoV-19/Scotland/QEUH-9F6FBC/2020|EPI_ISL_595650|2 ···················· ···················· ···················· ···················· ···················· ····················
M 1024 hCoV-19/Scotland/QEUH-9F6FDA/2020|EPI_ISL_595651|2 ···················· ···················· ···················· ···················· ···················· ····················
M 1025 hCoV-19/England/QEUH-A00E39/2020|EPI_ISL_584901|20 ···················· ···················· ···················· ···················· ···················· ····················
M 1026 hCoV-19/England/QEUH-A0114E/2020|EPI_ISL_584903|20 ···················· ···················· ···················· ···················· ···················· ····················
M 1027 hCoV-19/England/QEUH-A0115D/2020|EPI_ISL_584904|20 ···················· ···················· ···················· ···················· ···················· ····················
M 1028 hCoV-19/England/QEUH-A0116C/2020|EPI_ISL_584905|20 ···················· ···················· ···················· ···················· ···················· ····················
M 1030 hCoV-19/England/ALDP-A2FDEC/2020|EPI_ISL_587124|20 ···················· ···················· ···················· ···················· ···················· ····················
M 1031 hCoV-19/England/ALDP-A2FEF8/2020|EPI_ISL_587127|20 ···················· ···················· ···················· ···················· ···················· ····················
M 1032 hCoV-19/England/ALDP-A2F13B/2020|EPI_ISL_587128|20 ···················· ···················· ···················· ···················· ···················· ····················
M 1033 hCoV-19/England/ALDP-A2EB7A/2020|EPI_ISL_587129|20 ···················· ···················· ···················· ···················· ···················· ····················
M 1034 hCoV-19/England/ALDP-A2F0E3/2020|EPI_ISL_587131|20 ···················· ···················· ···················· ···················· ···················· ····················
M 1035 hCoV-19/England/ALDP-A2F7C0/2020|EPI_ISL_587132|20 ···················· ···················· ···················· ···················· ···················· ····················
M 1036 hCoV-19/England/ALDP-A2E98F/2020|EPI_ISL_587133|20 ···················· ···················· ···················· ···················· ···················· ····················
M 1037 hCoV-19/England/ALDP-A2EC68/2020|EPI_ISL_587134|20 ···················· ·A·················· ···················· ···················· ···················· ··A·················
M 1038 hCoV-19/England/ALDP-A2F669/2020|EPI_ISL_587135|20 ···················· ···················· ···················· ···················· ···················· ····················
M 1039 hCoV-19/England/ALDP-A2FB97/2020|EPI_ISL_587136|20 ···················· ···················· ···················· ···················· ···················· ····················
M 1040 hCoV-19/England/ALDP-A2FE8F/2020|EPI_ISL_587137|20 ···················· ···················· ···················· ···················· ···················· ····················
M 1041 hCoV-19/England/ALDP-A2FCEF/2020|EPI_ISL_587160|20 ···················· ···················· ···················· ···················· ···················· ····················
M 1042 hCoV-19/England/ALDP-A2FBB5/2020|EPI_ISL_587161|20 ···················· ···················· ···················· ···················· ···················· ····················
M 1043 hCoV-19/England/ALDP-A2F405/2020|EPI_ISL_587162|20 ···················· ···················· ···················· ···················· ···················· ····················
M 1044 hCoV-19/England/ALDP-A2EAAA/2020|EPI_ISL_587164|20 ···················· ···················· ···················· ···················· ···················· ····················
M 1045 hCoV-19/England/ALDP-A2F168/2020|EPI_ISL_587165|20 ···················· ···················· ···················· ···················· ···················· ····················
M 1046 hCoV-19/England/ALDP-A2F48D/2020|EPI_ISL_587166|20 ···················· ···················· ···················· ···················· ···················· ····················
M 1047 hCoV-19/England/ALDP-A2F0D4/2020|EPI_ISL_587167|20 ···················· ···················· ···················· ···················· ···················· ····················
M 1048 hCoV-19/England/ALDP-A2E8A0/2020|EPI_ISL_587168|20 ···················· ···················· ···················· ···················· ···················· ····················
M 1049 hCoV-19/England/ALDP-A2F8BE/2020|EPI_ISL_587169|20 ···················· ···················· ···················· ···················· ···················· ····················
M 1050 hCoV-19/England/ALDP-A2FC49/2020|EPI_ISL_587170|20 ···················· ···················· ···················· ···················· ···················· ····················
M 1051 hCoV-19/England/ALDP-A2F20B/2020|EPI_ISL_587171|20 ···················· ···················· ···················· ···················· ···················· ····················
M 1052 hCoV-19/England/ALDP-A2EFD8/2020|EPI_ISL_587172|20 ···················· ···················· ···················· ···················· ···················· ····················
M 1053 hCoV-19/England/ALDP-A2F766/2020|EPI_ISL_587173|20 ···················· ···················· ···················· ···················· ···················· ····················
M 1054 hCoV-19/England/ALDP-A2F1FF/2020|EPI_ISL_587174|20 ···················· ···················· ···················· ···················· ···················· ····················
M 1055 hCoV-19/England/ALDP-A2F177/2020|EPI_ISL_587175|20 ···················· ···················· ···················· ···················· ···················· ····················
M 1056 hCoV-19/England/ALDP-A2FD55/2020|EPI_ISL_587176|20 ···················· ···················· ···················· ···················· ···················· ····················
M 1057 hCoV-19/England/ALDP-A2E8FB/2020|EPI_ISL_587177|20 ···················· ···················· ···················· ···················· ···················· ····················
M 1058 hCoV-19/England/ALDP-A2EBE3/2020|EPI_ISL_587178|20 ···················· ···················· ···················· ···················· ···················· ····················
M 1059 hCoV-19/England/ALDP-A2F8CD/2020|EPI_ISL_587179|20 ···················· ···················· ···················· ···················· ···················· ····················
M 1060 hCoV-19/England/ALDP-A2F61E/2020|EPI_ISL_587180|20 ···················· ···················· ···················· ···················· ···················· ····················
M 1061 hCoV-19/England/ALDP-A2FDA0/2020|EPI_ISL_587181|20 ···················· ·A·················· ···················· ···················· ···················· ····················
M 1062 hCoV-19/England/ALDP-A2E846/2020|EPI_ISL_587182|20 ···················· ···················· ···················· ···················· ···················· ····················
M 1063 hCoV-19/England/ALDP-A2F229/2020|EPI_ISL_587183|20 ···················· ···················· ···················· ···················· ···················· ····················
M 1064 hCoV-19/England/ALDP-A2FE07/2020|EPI_ISL_587184|20 ···················· ···················· ···················· ···················· ···················· ····················
M 1065 hCoV-19/England/ALDP-A2F010/2020|EPI_ISL_587185|20 ···················· ···················· ···················· ···················· ···················· ····················
M 1066 hCoV-19/England/ALDP-A2FD19/2020|EPI_ISL_587186|20 ···················· ···················· ···················· ···················· ···················· ····················
M 1067 hCoV-19/England/ALDP-A2EC95/2020|EPI_ISL_587187|20 ···················· ···················· ···················· ···················· ···················· ····················
M 1068 hCoV-19/England/ALDP-A2ECA4/2020|EPI_ISL_587188|20 ···················· ···················· ···················· ···················· ···················· ····················

| | | | | | | | | |
|---|---|---|---|---|---|---|---|---|
| M | 1069 | hCoV-19/England/ALDP-A2FE43/2020\|EPI_ISL_587189\|20 | ···················· | ···················· | ···················· | ···················· | ···················· | ···················· |
| M | 1070 | hCoV-19/England/ALDP-A2F58A/2020\|EPI_ISL_587190\|20 | ···················· | ···················· | ···················· | ···················· | ···················· | ···················· |
| M | 1071 | hCoV-19/England/ALDP-A2F520/2020\|EPI_ISL_587191\|20 | ···················· | ···················· | ···················· | ···················· | ···················· | ···················· |
| M | 1072 | hCoV-19/England/ALDP-A2FC2B/2020\|EPI_ISL_587192\|20 | ···················· | ···················· | ···················· | ···················· | ···················· | ···················· |
| M | 1073 | hCoV-19/England/ALDP-A2FA21/2020\|EPI_ISL_587193\|20 | ···················· | ···················· | ···················· | ···················· | ···················· | ···················· |
| M | 1074 | hCoV-19/England/ALDP-A2FCFE/2020\|EPI_ISL_587194\|20 | ···················· | ···················· | ···················· | ···················· | ···················· | ···················· |
| M | 1075 | hCoV-19/England/ALDP-A2EC86/2020\|EPI_ISL_587195\|20 | ···················· | ···················· | ···················· | ···················· | ···················· | ···················· |
| M | 1076 | hCoV-19/England/ALDP-A2F64B/2020\|EPI_ISL_587196\|20 | ···················· | ···················· | ···················· | ···················· | ···················· | ···················· |
| M | 1077 | hCoV-19/England/ALDP-A2F70C/2020\|EPI_ISL_587197\|20 | ···················· | ···················· | ···················· | ···················· | ···················· | ···················· |
| M | 1078 | hCoV-19/England/ALDP-A2FB88/2020\|EPI_ISL_587198\|20 | ···················· | ···················· | ···················· | ···················· | ···················· | ···················· |
| M | 1079 | hCoV-19/England/ALDP-A2FEDA/2020\|EPI_ISL_587199\|20 | ···················· | ···················· | ···················· | ···················· | ···················· | ···················· |
| M | 1080 | hCoV-19/England/ALDP-A2FAC7/2020\|EPI_ISL_587215\|20 | ···················· | ···················· | ···················· | ···················· | ···················· | ···················· |
| M | 1081 | hCoV-19/England/ALDP-A2F256/2020\|EPI_ISL_587216\|20 | ···················· | ···················· | ···················· | ···················· | ···················· | ···················· |
| M | 1082 | hCoV-19/England/ALDP-A2EC4A/2020\|EPI_ISL_587217\|20 | ···················· | ···················· | ···················· | ···················· | ···················· | ···················· |
| M | 1083 | hCoV-19/England/ALDP-A2F687/2020\|EPI_ISL_587218\|20 | ···················· | ···················· | ···················· | ···················· | ···················· | ···················· |
| M | 1084 | hCoV-19/England/ALDP-A2F872/2020\|EPI_ISL_587219\|20 | ···················· | ···················· | ···················· | ···················· | ···················· | ···················· |
| M | 1085 | hCoV-19/England/ALDP-A2ECD1/2020\|EPI_ISL_587220\|20 | ···················· | ·A·················· | ···················· | ···················· | ···················· | ··A················· |
| M | 1086 | hCoV-19/England/ALDP-A2F21A/2020\|EPI_ISL_587221\|20 | ···················· | ···················· | ···················· | ···················· | ···················· | ···················· |
| M | 1087 | hCoV-19/England/ALDP-A2FA5E/2020\|EPI_ISL_587222\|20 | ···················· | ···················· | ···················· | ···················· | ···················· | ···················· |
| M | 1088 | hCoV-19/England/ALDP-A2F4AB/2020\|EPI_ISL_587223\|20 | ···················· | ···················· | ···················· | ···················· | ···················· | ···················· |
| M | 1089 | hCoV-19/England/ALDP-A2ECB3/2020\|EPI_ISL_587224\|20 | ···················· | ···················· | ···················· | ···················· | ···················· | ···················· |
| M | 1090 | hCoV-19/England/ALDP-A2ECE0/2020\|EPI_ISL_587225\|20 | ···················· | ···················· | ···················· | ···················· | ···················· | ···················· |
| M | 1091 | hCoV-19/England/ALDP-A2EE9F/2020\|EPI_ISL_587228\|20 | ···················· | ···················· | ···················· | ···················· | ···················· | ···················· |
| M | 1092 | hCoV-19/England/ALDP-A2F3EA/2020\|EPI_ISL_587229\|20 | ···················· | ···················· | ···················· | ···················· | ···················· | ···················· |
| M | 1093 | hCoV-19/England/QEUH-9F4DB4/2020\|EPI_ISL_588458\|20 | ···················· | ···················· | ···················· | ···················· | ···················· | ···················· |
| M | 1094 | hCoV-19/England/QEUH-9F4DA5/2020\|EPI_ISL_588459\|20 | ···················· | ·A·················· | ···················· | ···················· | ···················· | ··A················· |
| M | 1095 | hCoV-19/England/QEUH-9F47F2/2020\|EPI_ISL_588460\|20 | ···················· | ···················· | ···················· | ···················· | ···················· | ···················· |
| M | 1096 | hCoV-19/England/QEUH-9F5928/2020\|EPI_ISL_588461\|20 | ···················· | ···················· | ···················· | ···················· | ···················· | ···················· |
| M | 1097 | hCoV-19/England/QEUH-9F5867/2020\|EPI_ISL_588462\|20 | ···················· | ···················· | ···················· | ···················· | ···················· | ···················· |
| M | 1098 | hCoV-19/England/QEUH-9F4C5D/2020\|EPI_ISL_588463\|20 | ···················· | ···················· | ···················· | ···················· | ···················· | ···················· |
| M | 1099 | hCoV-19/England/QEUH-9F4BAB/2020\|EPI_ISL_588464\|20 | ···················· | ···················· | ···················· | ···················· | ···················· | ···················· |
| M | 1100 | hCoV-19/England/QEUH-9F51C6/2020\|EPI_ISL_588465\|20 | ···················· | ···················· | ···················· | ···················· | ···················· | ···················· |
| M | 1101 | hCoV-19/England/QEUH-9F4EB1/2020\|EPI_ISL_588466\|20 | ···················· | ···················· | ···················· | ···················· | ···················· | ···················· |
| M | 1102 | hCoV-19/England/QEUH-9F4D0F/2020\|EPI_ISL_588467\|20 | ···················· | ···················· | ···················· | ···················· | ···················· | ···················· |
| M | 1103 | hCoV-19/England/QEUH-9F5D95/2020\|EPI_ISL_588468\|20 | ···················· | ···················· | ···················· | ···················· | ···················· | ···················· |
| M | 1104 | hCoV-19/England/QEUH-9F409D/2020\|EPI_ISL_588490\|20 | ···················· | ···················· | ···················· | ···················· | ···················· | ···················· |
| M | 1105 | hCoV-19/England/QEUH-9F3298/2020\|EPI_ISL_588491\|20 | ···················· | ···················· | ···················· | ···················· | ···················· | ···················· |
| M | 1106 | hCoV-19/England/QEUH-9F3A54/2020\|EPI_ISL_588492\|20 | ···················· | ···················· | ···················· | ···················· | ···················· | ···················· |
| M | 1107 | hCoV-19/Northern Ireland/QEUH-9F3201/2020\|EPI_ISL_ | ···················· | ···················· | ···················· | ···················· | ···················· | ···················· |
| M | 1108 | hCoV-19/England/QEUH-9F2F65/2020\|EPI_ISL_588497\|20 | ···················· | ···················· | ···················· | ···················· | ···················· | ···················· |
| M | 1109 | hCoV-19/England/QEUH-9F369C/2020\|EPI_ISL_588498\|20 | ···················· | ·A·················· | ···················· | ···················· | ···················· | ··A················· |
| M | 1110 | hCoV-19/England/QEUH-9F36F6/2020\|EPI_ISL_588499\|20 | ···················· | ···················· | ···················· | ···················· | ···················· | ···················· |
| M | 1111 | hCoV-19/England/QEUH-9F31C8/2020\|EPI_ISL_588500\|20 | ···················· | ···················· | ···················· | ···················· | ···················· | ···················· |
| M | 1112 | hCoV-19/England/QEUH-9F3BD9/2020\|EPI_ISL_588501\|20 | ···················· | ···················· | ···················· | ···················· | ···················· | ···················· |
| M | 1113 | hCoV-19/England/QEUH-9F322F/2020\|EPI_ISL_588502\|20 | ···················· | ···················· | ···················· | ···················· | ···················· | ···················· |
| M | 1114 | hCoV-19/England/QEUH-9F3F37/2020\|EPI_ISL_588503\|20 | ···················· | ···················· | ···················· | ···················· | ···················· | ···················· |
| M | 1115 | hCoV-19/England/QEUH-9F2EB3/2020\|EPI_ISL_588504\|20 | ···················· | ···················· | ···················· | ···················· | ···················· | ···················· |
| M | 1116 | hCoV-19/England/QEUH-9F39EE/2020\|EPI_ISL_588505\|20 | ···················· | ···················· | ···················· | ···················· | ···················· | ···················· |
| M | 1117 | hCoV-19/England/QEUH-9F374E/2020\|EPI_ISL_588506\|20 | ···················· | ···················· | ···················· | ···················· | ···················· | ···················· |
| M | 1118 | hCoV-19/England/QEUH-9F35EA/2020\|EPI_ISL_588507\|20 | ···················· | ···················· | ···················· | ···················· | ···················· | ···················· |
| M | 1119 | hCoV-19/England/QEUH-9F42D3/2020\|EPI_ISL_588508\|20 | ···················· | ···················· | ···················· | ···················· | ···················· | ···················· |
| M | 1120 | hCoV-19/England/QEUH-9F3E49/2020\|EPI_ISL_588509\|20 | ···················· | ···················· | ···················· | ···················· | ···················· | ···················· |
| M | 1121 | hCoV-19/England/QEUH-9F3FFB/2020\|EPI_ISL_588510\|20 | ···················· | ···················· | ···················· | ···················· | ···················· | ···················· |
| M | 1122 | hCoV-19/England/QEUH-9F37E4/2020\|EPI_ISL_588511\|20 | ···················· | ···················· | ···················· | ···················· | ···················· | ···················· |
| M | 1123 | hCoV-19/England/QEUH-9F319B/2020\|EPI_ISL_588512\|20 | ···················· | ···················· | ···················· | ···················· | ···················· | ···················· |
| M | 1124 | hCoV-19/England/QEUH-9F36D8/2020\|EPI_ISL_588513\|20 | ···················· | ···················· | ···················· | ···················· | ···················· | ···················· |
| M | 1125 | hCoV-19/England/QEUH-9F382D/2020\|EPI_ISL_588514\|20 | ···················· | ···················· | ···················· | ···················· | ···················· | ···················· |
| M | 1126 | hCoV-19/England/QEUH-9F2FED/2020\|EPI_ISL_588515\|20 | ···················· | ···················· | ···················· | ···················· | ···················· | ···················· |
| M | 1127 | hCoV-19/England/QEUH-9F36AB/2020\|EPI_ISL_588516\|20 | ···················· | ···················· | ···················· | ···················· | ···················· | ···················· |
| M | 1128 | hCoV-19/England/QEUH-9F3C9A/2020\|EPI_ISL_588517\|20 | ···················· | ···················· | ···················· | ···················· | ···················· | ···················· |
| M | 1129 | hCoV-19/England/QEUH-9F3A18/2020\|EPI_ISL_588518\|20 | ···················· | ···················· | ···················· | ···················· | ···················· | ···················· |
| M | 1130 | hCoV-19/England/QEUH-9F54EB/2020\|EPI_ISL_588519\|20 | ···················· | ···················· | ···················· | ···················· | ···················· | ···················· |
| M | 1131 | hCoV-19/England/QEUH-9F3508/2020\|EPI_ISL_588520\|20 | ···················· | ···················· | ···················· | ···················· | ···················· | ···················· |
| M | 1132 | hCoV-19/England/QEUH-9F326B/2020\|EPI_ISL_588521\|20 | ···················· | ···················· | ···················· | ···················· | ···················· | ···················· |
| M | 1133 | hCoV-19/England/QEUH-9F4042/2020\|EPI_ISL_588522\|20 | ···················· | ···················· | ···················· | ···················· | ···················· | ···················· |
| M | 1134 | hCoV-19/England/QEUH-9F2CB9/2020\|EPI_ISL_588546\|20 | ···················· | ···················· | ···················· | ···················· | ···················· | ···················· |
| M | 1135 | hCoV-19/England/QEUH-9F3DC4/2020\|EPI_ISL_588547\|20 | ···················· | ···················· | ···················· | ···················· | ···················· | ···················· |
| M | 1136 | hCoV-19/England/QEUH-9F41A9/2020\|EPI_ISL_588548\|20 | ···················· | ···················· | ···················· | ···················· | ···················· | ···················· |
| M | 1137 | hCoV-19/England/QEUH-9F3B33/2020\|EPI_ISL_588549\|20 | ···················· | ···················· | ···················· | ···················· | ···················· | ···················· |
| M | 1138 | hCoV-19/England/QEUH-9F31D7/2020\|EPI_ISL_588550\|20 | ···················· | ···················· | ···················· | ···················· | ···················· | ···················· |
| M | 1139 | hCoV-19/England/QEUH-9F2E2C/2020\|EPI_ISL_588551\|20 | ···················· | ···················· | ···················· | ···················· | ···················· | ···················· |
| M | 1140 | hCoV-19/England/QEUH-9F54CD/2020\|EPI_ISL_588552\|20 | ···················· | ···················· | ···················· | ···················· | ···T················ | ···················· |
| M | 1141 | hCoV-19/England/QEUH-9F4121/2020\|EPI_ISL_588553\|20 | ···················· | ···················· | ···················· | ···················· | ···················· | ···················· |
| M | 1142 | hCoV-19/England/QEUH-9F366F/2020\|EPI_ISL_588555\|20 | ···················· | ···················· | ···················· | ···················· | ···················· | ···················· |
| M | 1143 | hCoV-19/England/QEUH-9F414F/2020\|EPI_ISL_588556\|20 | ···················· | ···················· | ···················· | ···················· | ···················· | ···················· |
| M | 1144 | hCoV-19/England/QEUH-9F33FF/2020\|EPI_ISL_588557\|20 | ···················· | ···················· | ···················· | ···················· | ···················· | ···················· |
| M | 1145 | hCoV-19/England/QEUH-9F3B42/2020\|EPI_ISL_588559\|20 | ···················· | ···················· | ···················· | ···················· | ···················· | ···················· |
| M | 1146 | hCoV-19/Wales/ALDP-9EA0B8/2020\|EPI_ISL_589780\|2020 | ···················· | ···················· | ···················· | ···················· | ···T················ | ···················· |
| M | 1147 | hCoV-19/Wales/MILK-9E2B19/2020\|EPI_ISL_589789\|2020 | ···················· | ···················· | ···················· | ···················· | ···················· | ···················· |
| M | 1148 | hCoV-19/England/QEUH-9DE58B/2020\|EPI_ISL_589790\|20 | ···················· | ···················· | ···················· | ···················· | ···················· | ···················· |

| | | | | | | | |
|---|---|---|---|---|---|---|---|
| M | 1149 hCoV-19/Wales/MILK-9E2BFB/2020\|EPI_ISL_589791\|2020 | ···················· | ···················· | ···················· | ···················· | ···················· | ···················· |
| M | 1150 hCoV-19/Wales/MILK-9E2E01/2020\|EPI_ISL_589792\|2020 | ···················· | ···················· | ···················· | ···················· | ···················· | ···················· |
| M | 1151 hCoV-19/Wales/MILK-9E1E02/2020\|EPI_ISL_589794\|2020 | ···················· | ···················· | ···················· | ···················· | ···················· | ···················· |
| M | 1152 hCoV-19/Wales/MILK-9E2E3E/2020\|EPI_ISL_589795\|2020 | ···················· | ···················· | ···················· | ···················· | ···················· | ···················· |
| M | 1153 hCoV-19/England/MILK-9E2214/2020\|EPI_ISL_589796\|20 | ···················· | ···················· | ···················· | ···················· | ···················· | ···················· |
| M | 1154 hCoV-19/Wales/MILK-9E0674/2020\|EPI_ISL_589797\|2020 | ···················· | ···················· | ···················· | ···················· | ···················· | ···················· |
| M | 1155 hCoV-19/Wales/MILK-9E0692/2020\|EPI_ISL_589798\|2020 | ···················· | ···················· | ···················· | ···················· | ···················· | ···················· |
| M | 1156 hCoV-19/Wales/MILK-9E05FF/2020\|EPI_ISL_589799\|2020 | ···················· | ···················· | ···················· | ···················· | ···················· | ···················· |
| M | 1157 hCoV-19/England/QEUH-9D09F6/2020\|EPI_ISL_589811\|20 | ···················· | ···················· | ···················· | ···················· | ···················· | ···················· |
| M | 1158 hCoV-19/England/QEUH-9D0167/2020\|EPI_ISL_589812\|20 | ···················· | ···················· | ···················· | ···················· | ···················· | ···················· |
| M | 1159 hCoV-19/England/QEUH-9D0DDC/2020\|EPI_ISL_589813\|20 | ···················· | ···················· | ···················· | ···················· | ···················· | ···················· |
| M | 1160 hCoV-19/England/QEUH-9D0765/2020\|EPI_ISL_589814\|20 | ···················· | ···················· | ···················· | ···················· | ···················· | ···················· |
| M | 1161 hCoV-19/England/QEUH-9CFE37/2020\|EPI_ISL_589815\|20 | ···················· | ···················· | ···················· | ···················· | ···················· | ···················· |
| M | 1162 hCoV-19/England/QEUH-9D0835/2020\|EPI_ISL_589816\|20 | ···················· | ···················· | ···················· | ···················· | ···················· | ···················· |
| M | 1163 hCoV-19/England/QEUH-9D00F1/2020\|EPI_ISL_589817\|20 | ···················· | ·A·················· | ···················· | ···················· | ···················· | ···················· |
| M | 1164 hCoV-19/England/QEUH-9CFCA6/2020\|EPI_ISL_589818\|20 | ···················· | ···················· | ···················· | ···················· | ···················· | ···················· |
| M | 1165 hCoV-19/England/QEUH-9D0695/2020\|EPI_ISL_589819\|20 | ···················· | ·A·················· | ···················· | ···················· | ···················· | ···················· |
| M | 1166 hCoV-19/England/QEUH-9CFDC1/2020\|EPI_ISL_589820\|20 | ···················· | ···················· | ···················· | ···················· | ···················· | ···················· |
| M | 1167 hCoV-19/England/QEUH-9D05B6/2020\|EPI_ISL_589821\|20 | ···················· | ···················· | ···················· | ···················· | ···················· | ···················· |
| M | 1168 hCoV-19/England/QEUH-9D04E6/2020\|EPI_ISL_589822\|20 | ···················· | ···················· | ···················· | ···················· | ···················· | ···················· |
| M | 1169 hCoV-19/England/QEUH-9CFD67/2020\|EPI_ISL_589823\|20 | ···················· | ···················· | ···················· | ···················· | ···················· | ···················· |
| M | 1170 hCoV-19/England/QEUH-9CFFE9/2020\|EPI_ISL_589824\|20 | ···················· | ···················· | ···················· | ···················· | ···················· | ···················· |
| M | 1171 hCoV-19/England/QEUH-9D01D0/2020\|EPI_ISL_589825\|20 | ···················· | ···················· | ···················· | ···················· | ···················· | ···················· |
| M | 1172 hCoV-19/England/QEUH-9CF9EB/2020\|EPI_ISL_589826\|20 | ···················· | ···················· | ···················· | ···················· | ···················· | ···················· |
| M | 1173 hCoV-19/England/QEUH-9CFFF8/2020\|EPI_ISL_589827\|20 | ···················· | ···················· | ···················· | ···················· | ···················· | ···················· |
| M | 1174 hCoV-19/England/QEUH-9D0F03/2020\|EPI_ISL_589828\|20 | ···················· | ···················· | ···················· | ···················· | ···················· | ···················· |
| M | 1175 hCoV-19/England/QEUH-9D04B9/2020\|EPI_ISL_589829\|20 | ···················· | ···················· | ···················· | ···················· | ···················· | ···················· |
| M | 1176 hCoV-19/England/QEUH-9D005B/2020\|EPI_ISL_589830\|20 | ···················· | ···················· | ···················· | ···················· | ···················· | ···················· |
| M | 1177 hCoV-19/England/QEUH-9D0C66/2020\|EPI_ISL_589831\|20 | ···················· | ···················· | ···················· | ···················· | ···················· | ···················· |
| M | 1178 hCoV-19/England/QEUH-9CF9DC/2020\|EPI_ISL_589832\|20 | ···················· | ···················· | ···················· | ···················· | ···················· | ···················· |
| M | 1179 hCoV-19/England/QEUH-9D0DFA/2020\|EPI_ISL_589833\|20 | ···················· | ···················· | ···················· | ···················· | ···················· | ···················· |
| M | 1180 hCoV-19/England/QEUH-9D0B5A/2020\|EPI_ISL_589834\|20 | ···················· | ···················· | ···················· | ···················· | ···················· | ···················· |
| M | 1181 hCoV-19/England/QEUH-9D0361/2020\|EPI_ISL_589836\|20 | ···················· | ···················· | ···················· | ···················· | ···················· | ···················· |
| M | 1182 hCoV-19/Wales/QEUH-9CFF70/2020\|EPI_ISL_589839\|2020 | ···················· | ···················· | ···················· | ···················· | ···················· | ···················· |
| M | 1183 hCoV-19/England/QEUH-9CFE64/2020\|EPI_ISL_589840\|20 | ···················· | ···················· | ···················· | ···················· | ···················· | ···················· |
| M | 1184 hCoV-19/England/QEUH-9D05A7/2020\|EPI_ISL_589841\|20 | ···················· | ·A·················· | ···················· | ···················· | ···················· | ···A················ |
| M | 1185 hCoV-19/England/QEUH-9D0B78/2020\|EPI_ISL_589842\|20 | ···················· | ···················· | ···················· | ···················· | ···················· | ···················· |
| M | 1186 hCoV-19/England/QEUH-9D0B3C/2020\|EPI_ISL_589843\|20 | ···················· | ···················· | ···················· | ···················· | ···················· | ···················· |
| M | 1187 hCoV-19/England/QEUH-9CFA8E/2020\|EPI_ISL_589844\|20 | ···················· | ···················· | ···················· | ···················· | ···················· | ···················· |
| M | 1188 hCoV-19/England/QEUH-9D0BB4/2020\|EPI_ISL_589845\|20 | ···················· | ···················· | ···················· | ···················· | ···················· | ···················· |
| M | 1189 hCoV-19/England/QEUH-9CFC00/2020\|EPI_ISL_589847\|20 | ···················· | ···················· | ···················· | ···················· | ···················· | ···················· |
| M | 1190 hCoV-19/England/QEUH-9CF9AF/2020\|EPI_ISL_589848\|20 | ···················· | ···················· | ···················· | ···················· | ···················· | ···················· |
| M | 1191 hCoV-19/England/QEUH-9CF8EE/2020\|EPI_ISL_589849\|20 | ···················· | ···················· | ···················· | ···················· | ···················· | ···················· |
| M | 1192 hCoV-19/England/QEUH-9D010D/2020\|EPI_ISL_589852\|20 | ···················· | ···················· | ···················· | ···················· | ···················· | ···················· |
| M | 1193 hCoV-19/England/QEUH-9D03BC/2020\|EPI_ISL_589853\|20 | ···················· | ·A·················· | ···················· | ···················· | ···················· | ···················· |
| M | 1194 hCoV-19/England/QEUH-9D0B2D/2020\|EPI_ISL_589854\|20 | ···················· | ·A·················· | ···················· | ···················· | ···················· | ···················· |
| M | 1195 hCoV-19/England/QEUH-9D02CE/2020\|EPI_ISL_589878\|20 | ···················· | ···················· | ···················· | ···················· | ···················· | ···················· |
| M | 1196 hCoV-19/England/QEUH-9D0F6D/2020\|EPI_ISL_589879\|20 | ···················· | ···················· | ···················· | ···················· | ······T············· | ···················· |
| M | 1197 hCoV-19/England/QEUH-9D0307/2020\|EPI_ISL_589880\|20 | ···················· | ···················· | ···················· | ···················· | ···················· | ···················· |
| M | 1198 hCoV-19/England/QEUH-9D0EAC/2020\|EPI_ISL_589881\|20 | ···················· | ···················· | ···················· | ···················· | ···················· | ···················· |
| M | 1199 hCoV-19/England/QEUH-9D07ED/2020\|EPI_ISL_589882\|20 | ···················· | ···················· | ···················· | ···················· | ···················· | ···················· |
| M | 1200 hCoV-19/England/QEUH-9D0325/2020\|EPI_ISL_589883\|20 | ···················· | ·A·················· | ···················· | ···················· | ···················· | ···················· |
| M | 1201 hCoV-19/England/QEUH-9D08DB/2020\|EPI_ISL_589884\|20 | ···················· | ···················· | ···················· | ···················· | ···················· | ···················· |
| M | 1202 hCoV-19/England/QEUH-9CFDB2/2020\|EPI_ISL_589885\|20 | ···················· | ···················· | ···················· | ···················· | ···················· | ···················· |
| M | 1203 hCoV-19/England/QEUH-9D0194/2020\|EPI_ISL_589886\|20 | ···················· | ···················· | ···················· | ···················· | ···················· | ···················· |
| M | 1204 hCoV-19/England/QEUH-9D001F/2020\|EPI_ISL_589887\|20 | ···················· | ···················· | ···················· | ···················· | ···················· | ···················· |
| M | 1205 hCoV-19/England/QEUH-9D06C2/2020\|EPI_ISL_589889\|20 | ···················· | ·A·················· | ···················· | ···················· | ···················· | ···A················ |
| M | 1206 hCoV-19/USA/MN-MDH-1760/2020\|EPI_ISL_590765\|2020-0 | ···················· | ···················· | ···················· | ···················· | ···················· | ···················· |
| M | 1207 hCoV-19/USA/MN-MDH-1761/2020\|EPI_ISL_590766\|2020-0 | ···················· | ···················· | ···················· | ···················· | ···················· | ···················· |
| M | 1208 hCoV-19/USA/MN-MDH-1762/2020\|EPI_ISL_590767\|2020-0 | ···················· | ···················· | ···················· | ···················· | ···················· | ···················· |
| M | 1209 hCoV-19/USA/MN-MDH-1763/2020\|EPI_ISL_590768\|2020-0 | ···················· | ···················· | ···················· | ···················· | ···················· | ···················· |
| M | 1210 hCoV-19/USA/MN-MDH-1764/2020\|EPI_ISL_590769\|2020-0 | ···················· | ···················· | ···················· | ···················· | ···················· | ···················· |
| M | 1211 hCoV-19/USA/MN-MDH-1766/2020\|EPI_ISL_590770\|2020-0 | ···················· | ···················· | ···················· | ···················· | ···················· | ···················· |
| M | 1212 hCoV-19/USA/MN-MDH-1767/2020\|EPI_ISL_590771\|2020-0 | ···················· | ···················· | ···················· | ···················· | ···················· | ···················· |
| M | 1213 hCoV-19/USA/MN-MDH-1768/2020\|EPI_ISL_590772\|2020-0 | ···················· | ···················· | ···················· | ···················· | ···················· | ···················· |
| M | 1214 hCoV-19/USA/MN-MDH-1769/2020\|EPI_ISL_590773\|2020-0 | ···················· | ···················· | ···················· | ···················· | ·····W·············· | ···················· |
| M | 1215 hCoV-19/USA/MN-MDH-1770/2020\|EPI_ISL_590774\|2020-0 | ···················· | ···················· | ···················· | ···················· | ···················· | ···················· |
| M | 1216 hCoV-19/USA/MN-MDH-1771/2020\|EPI_ISL_590775\|2020-0 | ···················· | ···················· | ···················· | ···················· | ···················· | ···················· |
| M | 1217 hCoV-19/USA/MN-MDH-1772/2020\|EPI_ISL_590776\|2020-0 | ···················· | ···················· | ···················· | ···················· | ···················· | ···················· |
| M | 1218 hCoV-19/USA/MN-MDH-1774/2020\|EPI_ISL_590777\|2020-0 | ···················· | ···················· | ···················· | ···················· | ···················· | ···················· |
| M | 1219 hCoV-19/USA/MN-MDH-1775/2020\|EPI_ISL_590778\|2020-0 | ···················· | ···················· | ···················· | ···················· | ···················· | ···················· |
| M | 1220 hCoV-19/USA/MN-MDH-1776/2020\|EPI_ISL_590779\|2020-0 | ···················· | ···················· | ···················· | ···················· | ···················· | ···················· |
| M | 1221 hCoV-19/USA/MN-MDH-1798/2020\|EPI_ISL_590800\|2020-0 | ···················· | ···················· | ···················· | ···················· | ···················· | ···················· |
| M | 1222 hCoV-19/USA/MN-MDH-1799/2020\|EPI_ISL_590801\|2020-0 | ···················· | ···················· | ···················· | ···················· | ···················· | ···················· |
| M | 1223 hCoV-19/USA/MN-MDH-1800/2020\|EPI_ISL_590802\|2020-0 | ···················· | ···················· | ···················· | ···················· | ···················· | ···················· |
| M | 1224 hCoV-19/USA/MN-MDH-1801/2020\|EPI_ISL_590803\|2020-0 | ···················· | ···················· | ···················· | ···················· | ···················· | ···················· |
| M | 1225 hCoV-19/USA/MN-MDH-1802/2020\|EPI_ISL_590804\|2020-0 | ···················· | ···················· | ···················· | ···················· | ···················· | ···················· |
| M | 1226 hCoV-19/USA/MN-MDH-1803/2020\|EPI_ISL_590805\|2020-0 | ···················· | ···················· | ···················· | ···················· | ···················· | ···················· |
| M | 1227 hCoV-19/USA/MN-MDH-1804/2020\|EPI_ISL_590806\|2020-0 | ···················· | ···················· | ···················· | ···················· | ···················· | ···················· |
| M | 1228 hCoV-19/USA/MN-MDH-1805/2020\|EPI_ISL_590807\|2020-0 | ···················· | ···················· | ···················· | ···················· | ···················· | ···················· |

| | | | | | | | | |
|---|---|---|---|---|---|---|---|---|
| M | 1229 | hCoV-19/USA/MN-MDH-1806/2020\|EPI_ISL_590808\|2020-0 | ···················· | ···················· | ···················· | ···················· | ···················· | ···················· |
| M | 1230 | hCoV-19/USA/MN-MDH-1807/2020\|EPI_ISL_590809\|2020-0 | ···················· | ···················· | ···················· | ···················· | ···················· | ···················· |
| M | 1231 | hCoV-19/USA/MN-MDH-1808/2020\|EPI_ISL_590810\|2020-0 | ···················· | ···················· | ···················· | ···················· | ···················· | ···················· |
| M | 1232 | hCoV-19/USA/MN-MDH-1809/2020\|EPI_ISL_590811\|2020-0 | ···················· | ···················· | ···················· | ···················· | ···················· | ···················· |
| M | 1233 | hCoV-19/USA/MN-MDH-1810/2020\|EPI_ISL_590812\|2020-0 | ···················· | ···················· | ···················· | ···················· | ···················· | ···················· |
| M | 1234 | hCoV-19/USA/MN-MDH-1811/2020\|EPI_ISL_590813\|2020-0 | ···················· | ···················· | ···················· | ···················· | ···················· | ···················· |
| M | 1235 | hCoV-19/USA/MN-MDH-1812/2020\|EPI_ISL_590814\|2020-0 | ···················· | ···················· | ···················· | ···················· | ···················· | ···················· |
| M | 1236 | hCoV-19/USA/MN-MDH-1813/2020\|EPI_ISL_590815\|2020-0 | ···················· | ···················· | ···················· | ···················· | ···················· | ···················· |
| M | 1237 | hCoV-19/USA/MN-MDH-1814/2020\|EPI_ISL_590816\|2020-0 | ···················· | ···················· | ···················· | ···················· | ···················· | ···················· |
| M | 1238 | hCoV-19/USA/MN-MDH-1815/2020\|EPI_ISL_590817\|2020-0 | ···················· | ···················· | ···················· | ···················· | ···················· | ···················· |
| M | 1239 | hCoV-19/USA/MN-MDH-1816/2020\|EPI_ISL_590818\|2020-0 | ···················· | ···················· | ···················· | ···················· | ···················· | ···················· |
| M | 1240 | hCoV-19/USA/MN-MDH-1817/2020\|EPI_ISL_590819\|2020-0 | ···················· | ···················· | ···················· | ···················· | ···················· | ···················· |
| M | 1241 | hCoV-19/USA/MN-MDH-1818/2020\|EPI_ISL_590820\|2020-0 | ···················· | ···················· | ···················· | ···················· | ···················· | ···················· |
| M | 1242 | hCoV-19/USA/MN-MDH-1819/2020\|EPI_ISL_590821\|2020-0 | ···················· | ···················· | ···················· | ···················· | ···················· | ···················· |
| M | 1243 | hCoV-19/USA/MN-MDH-1820/2020\|EPI_ISL_590822\|2020-0 | ···················· | ····G·····A········· | ···················· | ···················· | ···················· | ···················· |
| M | 1244 | hCoV-19/Switzerland/ZH-UZH-IMV1/2020\|EPI_ISL_59082 | ···················· | ···················· | ···················· | ···················· | ···················· | ···················· |
| M | 1245 | hCoV-19/Switzerland/ZH-UZH-IMV2/2020\|EPI_ISL_59082 | ···················· | ···················· | ···················· | ···················· | ···················· | ···················· |
| M | 1246 | hCoV-19/Switzerland/ZH-UZH-IMV3/2020\|EPI_ISL_59082 | ···················· | ···················· | ···················· | ···················· | ···················· | ···················· |
| M | 1247 | hCoV-19/Switzerland/ZH-UZH-IMV4/2020\|EPI_ISL_59082 | ···················· | ···················· | ···················· | ···················· | ···················· | ···················· |
| M | 1248 | hCoV-19/Switzerland/ZH-UZH-IMV5/2020\|EPI_ISL_59082 | ···················· | ···················· | ···················· | ···················· | ···················· | ···················· |
| M | 1249 | hCoV-19/Switzerland/ZH-UZH-IMV6/2020\|EPI_ISL_59082 | ···················· | ·A·················· | ···················· | ···················· | ···················· | ···················· |
| M | 1250 | hCoV-19/Switzerland/ZH-UZH-IMV7/2020\|EPI_ISL_59082 | ···················· | ···················· | ···················· | ···················· | ···················· | ···················· |
| M | 1251 | hCoV-19/Switzerland/ZH-UZH-IMV8/2020\|EPI_ISL_59083 | ···················· | ···················· | ···················· | ···················· | ···················· | ···················· |
| M | 1252 | hCoV-19/Switzerland/ZH-UZH-IMV9/2020\|EPI_ISL_59083 | ···················· | ···················· | ···················· | ···················· | ···················· | ···················· |
| M | 1253 | hCoV-19/Switzerland/TG-UZH-IMV10/2020\|EPI_ISL_5908 | ···················· | ···················· | ···················· | ···················· | ···················· | ···················· |
| M | 1254 | hCoV-19/Switzerland/AG-UZH-IMV11/2020\|EPI_ISL_5908 | ···················· | ·A·················· | ···················· | ···················· | ···················· | ···················· |
| M | 1255 | hCoV-19/Switzerland/AG-UZH-IMV12/2020\|EPI_ISL_5908 | ···················· | ···················· | ···················· | ···················· | ···················· | ···················· |
| M | 1256 | hCoV-19/Switzerland/AG-UZH-IMV13/2020\|EPI_ISL_5908 | ···················· | ···················· | ···················· | ···················· | ···················· | ···················· |
| M | 1257 | hCoV-19/Switzerland/ZH-UZH-IMV14/2020\|EPI_ISL_5908 | ···················· | ···················· | ···················· | ···················· | ···················· | ···················· |
| M | 1258 | hCoV-19/USA/MN-MDH-1777/2020\|EPI_ISL_590837\|2020-0 | ···················· | ···················· | ···················· | ···················· | ···················· | ···················· |
| M | 1259 | hCoV-19/Australia/VIC14955/2020\|EPI_ISL_593078\|202 | ···················· | ···················· | ···················· | ···················· | ···················· | ···················· |
| M | 1260 | hCoV-19/Australia/VIC14956/2020\|EPI_ISL_593079\|202 | ···················· | ···················· | ···················· | ···················· | ···················· | ···················· |
| M | 1263 | hCoV-19/Australia/VIC14959/2020\|EPI_ISL_593082\|202 | ···················· | ···················· | ···················· | ···················· | ···················· | ···················· |
| M | 1264 | hCoV-19/Australia/VIC14960/2020\|EPI_ISL_593083\|202 | ···················· | ···················· | ···················· | ···················· | ···················· | ···················· |
| M | 1265 | hCoV-19/Australia/VIC14963/2020\|EPI_ISL_593084\|202 | ···················· | ···················· | ···················· | ···················· | ···················· | ···················· |
| M | 1266 | hCoV-19/Australia/VIC14965/2020\|EPI_ISL_593085\|202 | ···················· | ···················· | ···················· | ···················· | ···················· | ···················· |
| M | 1267 | hCoV-19/Australia/VIC14972/2020\|EPI_ISL_593086\|202 | ···················· | ···················· | ···················· | ···················· | ···················· | ···················· |
| M | 1268 | hCoV-19/Australia/VIC14975/2020\|EPI_ISL_593087\|202 | ···················· | ···················· | ···················· | ···················· | ···················· | ···················· |
| M | 1269 | hCoV-19/Australia/VIC14977/2020\|EPI_ISL_593088\|202 | ···················· | ···················· | ···················· | ···················· | ···················· | ···················· |
| M | 1271 | hCoV-19/Australia/VIC14982/2020\|EPI_ISL_593090\|202 | ···················· | ···················· | ···················· | ···················· | ···················· | ···················· |
| M | 1272 | hCoV-19/Australia/VIC14983/2020\|EPI_ISL_593091\|202 | ···················· | ···················· | ···················· | ···················· | ···················· | ···················· |
| M | 1273 | hCoV-19/Australia/VIC14984/2020\|EPI_ISL_593092\|202 | ···················· | ···················· | ···················· | ···················· | ···················· | ···················· |
| M | 1275 | hCoV-19/Australia/VIC14989/2020\|EPI_ISL_593094\|202 | ···················· | ···················· | ···················· | ···················· | ···················· | ···················· |
| M | 1277 | hCoV-19/Australia/VIC14994/2020\|EPI_ISL_593096\|202 | ···················· | ···················· | ···················· | ···················· | ···················· | ···················· |
| M | 1280 | hCoV-19/Australia/VIC15000/2020\|EPI_ISL_593099\|202 | ···················· | ···················· | ···················· | ···················· | ···················· | ···················· |
| M | 1281 | hCoV-19/Austria/CeMM0730/2020\|EPI_ISL_583673\|2020- | ···················· | ···················· | ···················· | ···················· | ···················· | ···················· |
| M | 1282 | hCoV-19/Austria/CeMM0731/2020\|EPI_ISL_583674\|2020- | ···················· | ···················· | ···················· | ···················· | ···················· | ···················· |
| M | 1283 | hCoV-19/Austria/CeMM0732/2020\|EPI_ISL_583675\|2020- | ···················· | ···················· | ···················· | ···················· | ···················· | ···················· |
| M | 1284 | hCoV-19/Austria/CeMM0734/2020\|EPI_ISL_583676\|2020- | ···················· | ···················· | ···················· | ···················· | ···················· | ···················· |
| M | 1285 | hCoV-19/Austria/CeMM0739/2020\|EPI_ISL_583678\|2020- | ···················· | ···················· | ···················· | ···················· | ···················· | ···················· |
| M | 1286 | hCoV-19/Austria/CeMM0740/2020\|EPI_ISL_583679\|2020- | ···················· | ···················· | ···················· | ···················· | ···················· | ···················· |
| M | 1287 | hCoV-19/Austria/CeMM0742/2020\|EPI_ISL_583680\|2020- | ···················· | ···················· | ···················· | ···················· | ···················· | ···················· |
| M | 1288 | hCoV-19/Austria/CeMM0743/2020\|EPI_ISL_583681\|2020- | ···················· | ···················· | ···················· | ···················· | ···················· | ···················· |
| M | 1290 | hCoV-19/Austria/CeMM0746/2020\|EPI_ISL_583683\|2020- | ···················· | ···················· | ···················· | ···················· | ···················· | ···················· |
| M | 1291 | hCoV-19/Austria/CeMM0747/2020\|EPI_ISL_583684\|2020- | ···················· | ···················· | ···················· | ···················· | ···················· | ···················· |
| M | 1292 | hCoV-19/Austria/CeMM0749/2020\|EPI_ISL_583685\|2020- | ···················· | ···················· | ···················· | ···················· | ···················· | ···················· |
| M | 1293 | hCoV-19/Austria/CeMM0750/2020\|EPI_ISL_583686\|2020- | ···················· | ···················· | ···················· | ···················· | ···················· | ···················· |
| M | 1294 | hCoV-19/Austria/CeMM0752/2020\|EPI_ISL_583687\|2020- | ···················· | ···················· | ···················· | ···················· | ···················· | ···················· |
| M | 1295 | hCoV-19/Austria/CeMM0753/2020\|EPI_ISL_583688\|2020- | ···················· | ···················· | ···················· | ···················· | ···················· | ···················· |
| M | 1296 | hCoV-19/Austria/CeMM0755/2020\|EPI_ISL_583689\|2020- | ···················· | ···················· | ···················· | ···················· | ···················· | ···················· |
| M | 1297 | hCoV-19/Austria/CeMM0756/2020\|EPI_ISL_583690\|2020- | ···················· | ···················· | ···················· | ···················· | ···················· | ···················· |
| M | 1298 | hCoV-19/Austria/CeMM0758/2020\|EPI_ISL_583691\|2020- | ···················· | ···················· | ···················· | ···················· | ···················· | ···················· |
| M | 1299 | hCoV-19/Austria/CeMM0767/2020\|EPI_ISL_583692\|2020- | ···················· | ···················· | ···················· | ···················· | ···················· | ···················· |
| M | 1300 | hCoV-19/Austria/CeMM0768/2020\|EPI_ISL_583693\|2020- | ···················· | ···················· | ···················· | ···················· | ···················· | ···················· |
| M | 1301 | hCoV-19/Austria/CeMM0775/2020\|EPI_ISL_583694\|2020- | ···················· | ···················· | ···················· | ···················· | ···················· | ···················· |
| M | 1302 | hCoV-19/Austria/CeMM0779/2020\|EPI_ISL_583696\|2020- | ···················· | ···················· | ···················· | ···················· | ···················· | ···················· |
| M | 1303 | hCoV-19/Austria/CeMM0780/2020\|EPI_ISL_583697\|2020- | ···················· | ···················· | ···················· | ···················· | ···················· | ···················· |
| M | 1304 | hCoV-19/Austria/CeMM0781/2020\|EPI_ISL_583698\|2020- | ···················· | ···················· | ···················· | ···················· | ···················· | ···················· |
| M | 1305 | hCoV-19/England/ALDP-A2F63C/2020\|EPI_ISL_587200\|20 | ···················· | ···················· | ···················· | ···················· | ···················· | ···················· |
| M | 1306 | hCoV-19/England/ALDP-A2EF6F/2020\|EPI_ISL_587201\|20 | ···················· | ···················· | ···················· | ···················· | ···················· | ···················· |
| M | 1307 | hCoV-19/England/ALDP-A2F71B/2020\|EPI_ISL_587202\|20 | ···················· | ···················· | ···················· | ···················· | ···················· | ···················· |
| M | 1308 | hCoV-19/England/ALDP-A2F292/2020\|EPI_ISL_587203\|20 | ···················· | ···················· | ···················· | ···················· | ···················· | ···················· |
| M | 1309 | hCoV-19/England/ALDP-A2F450/2020\|EPI_ISL_587204\|20 | ···················· | ···················· | ···················· | ···················· | ···················· | ···················· |
| M | 1310 | hCoV-19/England/ALDP-A2F3F9/2020\|EPI_ISL_587205\|20 | ···················· | ···················· | ···················· | ···················· | ···················· | ···················· |
| M | 1311 | hCoV-19/England/ALDP-A2F6E1/2020\|EPI_ISL_587206\|20 | ···················· | ···················· | ···················· | ···················· | ···················· | ···················· |
| M | 1312 | hCoV-19/England/ALDP-A2EDED/2020\|EPI_ISL_587207\|20 | ···················· | ···················· | ···················· | ···················· | ···················· | ···················· |
| M | 1313 | hCoV-19/England/ALDP-A2F65A/2020\|EPI_ISL_587208\|20 | ···················· | ···················· | ···················· | ···················· | ···················· | ···················· |
| M | 1314 | hCoV-19/England/ALDP-A2EA04/2020\|EPI_ISL_587209\|20 | ···················· | ···················· | ···················· | ···················· | ···················· | ···················· |
| M | 1315 | hCoV-19/England/ALDP-A2F0A7/2020\|EPI_ISL_587210\|20 | ···················· | ···················· | ···················· | ···················· | ······T············· | ···················· |
| M | 1316 | hCoV-19/England/ALDP-A2ECC2/2020\|EPI_ISL_587211\|20 | ···················· | ···················· | ···················· | ···················· | ···················· | ···················· |

```
M 1317 hCoV-19/England/ALDP-A2F960/2020|EPI_ISL_587212|20 ···················· ···················· ···················· ···················· ···················· ····················
M 1318 hCoV-19/England/ALDP-A2E943/2020|EPI_ISL_587213|20 ···················· ···················· ···················· ···················· ···················· ····················
M 1319 hCoV-19/England/ALDP-A2EDFC/2020|EPI_ISL_587214|20 ···················· ···················· ···················· ···················· ···················· ····················
M 1320 hCoV-19/England/ALDP-A2FE70/2020|EPI_ISL_587226|20 ···················· ···················· ···················· ···················· ···················· ····················
M 1321 hCoV-19/England/ALDP-A2FA30/2020|EPI_ISL_587227|20 ···················· ···················· ···················· ···················· ···················· ····················
M 1322 hCoV-19/England/ALDP-A2F3CC/2020|EPI_ISL_587230|20 ···················· ···················· ···················· ···················· ···················· ····················
M 1323 hCoV-19/England/ALDP-A2E882/2020|EPI_ISL_587231|20 ···················· ···················· ···················· ···················· ···················· ····················
M 1324 hCoV-19/England/ALDP-A2E794/2020|EPI_ISL_587232|20 ···················· ···················· ···················· ···················· ···················· ····················
M 1325 hCoV-19/England/ALDP-A2E8EC/2020|EPI_ISL_587233|20 ···················· ···················· ···················· ···················· ···················· ····················
M 1326 hCoV-19/England/ALDP-A2F678/2020|EPI_ISL_587234|20 ···················· ···················· ···················· ···················· ···················· ····················
M 1327 hCoV-19/England/ALDP-A2EB4D/2020|EPI_ISL_587236|20 ···················· ···················· ···················· ···················· ···················· ····················
M 1328 hCoV-19/England/QEUH-9F35AE/2020|EPI_ISL_588524|20 ···················· ···················· ···················· ···················· ····T··············· ····················
M 1329 hCoV-19/England/QEUH-9F3FEC/2020|EPI_ISL_588525|20 ···················· ···················· ···················· ···················· ···················· ····················
M 1330 hCoV-19/England/QEUH-9F3CA9/2020|EPI_ISL_588526|20 ···················· ···················· ···················· ···················· ···················· ····················
M 1331 hCoV-19/England/QEUH-9F39FD/2020|EPI_ISL_588527|20 ···················· ···················· ···················· ···················· ···················· ····················
M 1332 hCoV-19/England/QEUH-9F3BBB/2020|EPI_ISL_588528|20 ···················· ···················· ···················· ···················· ···················· ····················
M 1333 hCoV-19/England/QEUH-9F380F/2020|EPI_ISL_588529|20 ···················· ···················· ···················· ···················· ···················· ····················
M 1334 hCoV-19/England/QEUH-9F3C30/2020|EPI_ISL_588530|20 ···················· ···················· ···················· ···················· ···················· ····················
M 1335 hCoV-19/England/QEUH-9F34A1/2020|EPI_ISL_588531|20 ···················· ···················· ···················· ···················· ···················· ····················
M 1336 hCoV-19/England/QEUH-9F3D97/2020|EPI_ISL_588532|20 ···················· ···················· ···················· ···················· ···················· ····················
M 1337 hCoV-19/England/QEUH-9F3EC1/2020|EPI_ISL_588533|20 ···················· ···················· ···················· ···················· ···················· ····················
M 1338 hCoV-19/England/QEUH-9F3DA6/2020|EPI_ISL_588534|20 ···················· ···················· ···················· ···················· ···················· ····················
M 1339 hCoV-19/England/QEUH-9F40D9/2020|EPI_ISL_588535|20 ···················· ···················· ···················· ···················· ···················· ····················
M 1340 hCoV-19/England/QEUH-9F35F9/2020|EPI_ISL_588536|20 ···················· ···················· ···················· ···················· ···················· ····················
M 1341 hCoV-19/England/QEUH-9F408E/2020|EPI_ISL_588537|20 ···················· ···················· ···················· ···················· ···················· ····················
M 1342 hCoV-19/England/QEUH-9F2EE0/2020|EPI_ISL_588538|20 ···················· ···················· ···················· ···················· ···················· ····················
M 1343 hCoV-19/England/QEUH-9F40CA/2020|EPI_ISL_588539|20 ···················· ···················· ···················· ···················· ···················· ····················
M 1344 hCoV-19/England/QEUH-9F3E76/2020|EPI_ISL_588540|20 ···················· ···················· ···················· ···················· ···················· ····················
M 1345 hCoV-19/England/QEUH-9F558E/2020|EPI_ISL_588542|20 ···················· ···················· ···················· ···················· ···················· ····················
M 1346 hCoV-19/England/QEUH-9F332C/2020|EPI_ISL_588543|20 ···················· ···················· ···················· ···················· ···················· ····················
M 1347 hCoV-19/England/QEUH-9F359F/2020|EPI_ISL_588544|20 ···················· ···················· ···················· ···················· ···················· ····················
M 1348 hCoV-19/Wales/QEUH-9F32C5/2020|EPI_ISL_588545|2020 ···················· ···················· ···················· ···················· ···················· ····················
M 1349 hCoV-19/England/QEUH-9F4385/2020|EPI_ISL_588558|20 ···················· ···················· ···················· ···················· ···················· ····················
M 1350 hCoV-19/England/QEUH-9F39C0/2020|EPI_ISL_588560|20 ···················· ···················· ···················· ···················· ···················· ····················
M 1351 hCoV-19/England/QEUH-9F3571/2020|EPI_ISL_588561|20 ···················· ···················· ···················· ···················· ···················· ····················
M 1352 hCoV-19/England/QEUH-9F38D2/2020|EPI_ISL_588562|20 ···················· ···················· ···················· ···················· ···················· ····················
M 1353 hCoV-19/England/QEUH-9F415E/2020|EPI_ISL_588563|20 ···················· ···················· ···················· ···················· ······T············· ····················
M 1354 hCoV-19/England/QEUH-9F30E9/2020|EPI_ISL_588564|20 ···················· ···················· ···················· ···················· ···················· ····················
M 1355 hCoV-19/England/QEUH-9F3474/2020|EPI_ISL_588565|20 ···················· ···················· ···················· ···················· ···················· ····················
M 1356 hCoV-19/England/QEUH-9F37F3/2020|EPI_ISL_588566|20 ···················· ···················· ···················· ···················· ···················· ····················
M 1357 hCoV-19/England/QEUH-9F3E3A/2020|EPI_ISL_588567|20 ···················· ···················· ···················· ···················· ···················· ····················
M 1358 hCoV-19/Wales/MILK-9D4C26/2020|EPI_ISL_589800|2020 ···················· ···················· ···················· ···················· ···················· ····················
M 1359 hCoV-19/Wales/MILK-9D3A5A/2020|EPI_ISL_589801|2020 ···················· ···················· ···················· ···················· ···················· ····················
M 1360 hCoV-19/Wales/ALDP-9CBBAD/2020|EPI_ISL_589802|2020 ···················· ···················· ···················· ···················· ···················· ····················
M 1361 hCoV-19/England/QEUH-9CFD58/2020|EPI_ISL_589804|20 ···················· ···················· ···················· ···················· ···················· ····················
M 1362 hCoV-19/England/QEUH-9CFABB/2020|EPI_ISL_589805|20 ···················· ···················· ···················· ···················· ···················· ····················
M 1363 hCoV-19/England/QEUH-9CFDD0/2020|EPI_ISL_589806|20 ···················· ·A·················· ···················· ···················· ···················· ····················
M 1364 hCoV-19/England/QEUH-9CFC5B/2020|EPI_ISL_589807|20 ···················· ···················· ···················· ···················· ···················· ····················
M 1365 hCoV-19/England/QEUH-9D0343/2020|EPI_ISL_589808|20 ···················· ···················· ···················· ···················· ···················· ····················
M 1366 hCoV-19/England/QEUH-9D012B/2020|EPI_ISL_589809|20 ···················· ···················· ···················· ···················· ···················· ····················
M 1367 hCoV-19/England/QEUH-9D0DBE/2020|EPI_ISL_589810|20 ···················· ···················· ···················· ···················· ···················· ····················
M 1368 hCoV-19/England/QEUH-9D0A5D/2020|EPI_ISL_589855|20 ···················· ···················· ···················· ···················· ···················· ····················
M 1369 hCoV-19/England/QEUH-9D052F/2020|EPI_ISL_589856|20 ···················· ···················· ···················· ···················· ···················· ····················
M 1370 hCoV-19/England/QEUH-9D0316/2020|EPI_ISL_589859|20 ···················· ···················· ···················· ···················· ···················· ····················
M 1371 hCoV-19/England/QEUH-9D006A/2020|EPI_ISL_589860|20 ···················· ···················· ···················· ···················· ···················· ····················
M 1372 hCoV-19/England/QEUH-9D0D09/2020|EPI_ISL_589861|20 ···················· ···················· ···················· ···················· ······T············· ····················
M 1373 hCoV-19/Wales/QEUH-9CFFDA/2020|EPI_ISL_589862|2020 ···················· ···················· ···················· ···················· ···················· ····················
M 1374 hCoV-19/England/QEUH-9D1193/2020|EPI_ISL_589863|20 ···················· ···················· ···················· ···················· ···················· ····················
M 1375 hCoV-19/England/QEUH-9CFA33/2020|EPI_ISL_589864|20 ···················· ···················· ···················· ···················· ···················· ····················
M 1376 hCoV-19/England/QEUH-9D060E/2020|EPI_ISL_589865|20 ···················· ···················· ···················· ···················· ···················· ····················
M 1377 hCoV-19/England/QEUH-9CFC6A/2020|EPI_ISL_589866|20 ···················· ···················· ···················· ···················· ···················· ····················
M 1378 hCoV-19/England/QEUH-9CFE55/2020|EPI_ISL_589867|20 ···················· ···················· ···················· ···················· ···················· ····················
M 1379 hCoV-19/England/QEUH-9D0088/2020|EPI_ISL_589868|20 ···················· ·A·················· ···················· ···················· ···················· ····················
M 1380 hCoV-19/England/QEUH-9D0000/2020|EPI_ISL_589869|20 ···················· ···················· ···················· ···················· ···················· ····················
M 1381 hCoV-19/England/QEUH-9D0880/2020|EPI_ISL_589870|20 ···················· ···················· ···················· ···················· ···················· ····················
M 1382 hCoV-19/England/QEUH-9D0FD6/2020|EPI_ISL_589871|20 ···················· ···················· ···················· ···················· ···················· ····················
M 1383 hCoV-19/England/QEUH-9D089F/2020|EPI_ISL_589873|20 ···················· ·A·················· ···················· ···················· ···················· ····················
M 1384 hCoV-19/England/QEUH-9D0756/2020|EPI_ISL_589874|20 ···················· ···················· ···················· ···················· ···················· ····················
M 1385 hCoV-19/England/QEUH-9D0747/2020|EPI_ISL_589875|20 ···················· ···················· ···················· ···················· ······T············· ····················
M 1386 hCoV-19/England/QEUH-9CFBC7/2020|EPI_ISL_589876|20 ···················· ···················· ···················· ···················· ···················· ····················
M 1387 hCoV-19/England/QEUH-9D02EC/2020|EPI_ISL_589888|20 ···················· ···················· ···················· ···················· ···················· ····················
M 1388 hCoV-19/England/QEUH-9D0C48/2020|EPI_ISL_589890|20 ···················· ···················· ···················· ···················· ····T··············· ····················
M 1389 hCoV-19/England/QEUH-9D0B0F/2020|EPI_ISL_589893|20 ···················· ···················· ···················· ···················· ···················· ····················
M 1390 hCoV-19/England/QEUH-9CFE73/2020|EPI_ISL_589894|20 ···················· ···················· ···················· ···················· ···················· ····················
M 1391 hCoV-19/England/QEUH-9CFD94/2020|EPI_ISL_589897|20 ···················· ···················· ···················· ···················· ···················· ····················
M 1392 hCoV-19/England/QEUH-9D0E7F/2020|EPI_ISL_589898|20 ···················· ···················· ···················· ···················· ···················· ····················
M 1393 hCoV-19/Norway/4016/2020|EPI_ISL_590877|2020-09-28 ···················· ···················· ···················· ···················· ···················· ····················
M 1394 hCoV-19/Norway/3293/2020|EPI_ISL_590878|2020-09-05 ···················· ···················· ···················· ···················· ···················· ····················
M 1395 hCoV-19/Norway/3326/2020|EPI_ISL_590879|2020-09-04 ···················· ···················· ···················· ···················· ···················· ····················
M 1396 hCoV-19/Australia/VIC14789/2020|EPI_ISL_593000|202 ···················· ···················· ···················· ···················· ·······S············ ····················
```

| | | | | | | | |
|---|---|---|---|---|---|---|---|
| M | 1399 hCoV-19/Australia/VIC14799/2020\|EPI_ISL_593003\|202 | ···················· | ···················· | ···················· | ···················· | ···················· | ···················· |
| M | 1404 hCoV-19/Australia/VIC14814/2020\|EPI_ISL_593008\|202 | ···················· | ···················· | ···················· | ···················· | ···················· | ···················· |
| M | 1405 hCoV-19/Australia/VIC14815/2020\|EPI_ISL_593009\|202 | ···················· | ···················· | ···················· | ···················· | ···················· | ···················· |
| M | 1406 hCoV-19/Australia/VIC14816/2020\|EPI_ISL_593010\|202 | ···················· | ···················· | ···················· | ···················· | ···················· | ···················· |
| M | 1407 hCoV-19/Australia/VIC14820/2020\|EPI_ISL_593011\|202 | ···················· | ···················· | ···················· | ···················· | ···················· | ···················· |
| M | 1408 hCoV-19/Australia/VIC14821/2020\|EPI_ISL_593012\|202 | ···················· | ···················· | ···················· | ···················· | ···················· | ···················· |
| M | 1409 hCoV-19/Australia/VIC14822/2020\|EPI_ISL_593013\|202 | ···················· | ···················· | ···················· | ···················· | ···················· | ···················· |
| M | 1410 hCoV-19/Australia/VIC14825/2020\|EPI_ISL_593014\|202 | ···················· | ···················· | ···················· | ···················· | ···················· | ···················· |
| M | 1412 hCoV-19/Australia/VIC14827/2020\|EPI_ISL_593016\|202 | ···················· | ···················· | ···················· | ···················· | ···················· | ···················· |
| M | 1419 hCoV-19/Australia/VIC14839/2020\|EPI_ISL_593023\|202 | ···················· | ···················· | ···················· | ···················· | ···················· | ···················· |
| M | 1420 hCoV-19/Australia/VIC14841/2020\|EPI_ISL_593024\|202 | ···················· | ···················· | ···················· | ···················· | ···················· | ···················· |
| M | 1421 hCoV-19/Australia/VIC14842/2020\|EPI_ISL_593025\|202 | ···················· | ···················· | ···················· | ···················· | ···················· | ···················· |
| M | 1423 hCoV-19/Australia/VIC14844/2020\|EPI_ISL_593027\|202 | ···················· | ···················· | ···················· | ···················· | ···················· | ···················· |
| M | 1424 hCoV-19/Australia/VIC14846/2020\|EPI_ISL_593028\|202 | ···················· | ···················· | ···················· | ···················· | ···················· | ···················· |
| M | 1425 hCoV-19/Australia/VIC14847/2020\|EPI_ISL_593029\|202 | ···················· | ···················· | ···················· | ···················· | ···················· | ···················· |
| M | 1427 hCoV-19/Australia/VIC14850/2020\|EPI_ISL_593031\|202 | ···················· | ···················· | ···················· | ···················· | ···················· | ···················· |
| M | 1428 hCoV-19/Australia/VIC14854/2020\|EPI_ISL_593032\|202 | ···················· | ···················· | ···················· | ···················· | ···················· | ···················· |
| M | 1430 hCoV-19/Australia/VIC14874/2020\|EPI_ISL_593040\|202 | ···················· | ···················· | ···················· | ···················· | ···················· | ···················· |
| M | 1432 hCoV-19/Australia/VIC14879/2020\|EPI_ISL_593042\|202 | ···················· | ···················· | ···················· | ···················· | ···················· | ···················· |
| M | 1433 hCoV-19/Australia/VIC14884/2020\|EPI_ISL_593043\|202 | ···················· | ···················· | ···················· | ···················· | ···················· | ···················· |
| M | 1435 hCoV-19/Australia/VIC14909/2020\|EPI_ISL_593056\|202 | ···················· | ···················· | ···················· | ···················· | ···················· | ···················· |
| M | 1436 hCoV-19/Australia/VIC14910/2020\|EPI_ISL_593057\|202 | ···················· | ···················· | ···················· | ···················· | ···················· | ···················· |
| M | 1437 hCoV-19/Australia/VIC14911/2020\|EPI_ISL_593058\|202 | ···················· | ···················· | ···················· | ···················· | ···················· | ···················· |
| M | 1438 hCoV-19/Australia/VIC14912/2020\|EPI_ISL_593059\|202 | ···················· | ···················· | ···················· | ···················· | ····K··············· | ···················· |
| M | 1439 hCoV-19/Australia/VIC14920/2020\|EPI_ISL_593060\|202 | ···················· | ···················· | ···················· | ···················· | ···················· | ···················· |
| M | 1441 hCoV-19/Australia/VIC14925/2020\|EPI_ISL_593062\|202 | ···················· | ···················· | ···················· | ···················· | ···················· | ···················· |
| M | 1443 hCoV-19/Australia/VIC14927/2020\|EPI_ISL_593064\|202 | ···················· | ···················· | ···················· | ···················· | ···················· | ···················· |
| M | 1444 hCoV-19/Australia/VIC14928/2020\|EPI_ISL_593065\|202 | ···················· | ···················· | ···················· | ···················· | ···················· | ···················· |
| M | 1445 hCoV-19/Australia/VIC14931/2020\|EPI_ISL_593066\|202 | ···················· | ···················· | ···················· | ···················· | ···················· | ···················· |
| M | 1446 hCoV-19/Australia/VIC14932/2020\|EPI_ISL_593067\|202 | ···················· | ···················· | ···················· | ···················· | ···················· | ···················· |
| M | 1452 hCoV-19/Australia/VIC14945/2020\|EPI_ISL_593073\|202 | ···················· | ···················· | ···················· | ···················· | ···················· | ···················· |
| M | 1453 hCoV-19/Australia/VIC14947/2020\|EPI_ISL_593074\|202 | ···················· | ···················· | ···················· | ···················· | ···················· | ···················· |
| M | 1454 hCoV-19/Australia/VIC14950/2020\|EPI_ISL_593075\|202 | ···················· | ···················· | ···················· | ···················· | ···················· | ···················· |
| M | 1455 hCoV-19/Australia/VIC14951/2020\|EPI_ISL_593076\|202 | ···················· | ···················· | ···················· | ···················· | ···················· | ···················· |
| M | 1457 hCoV-19/USA/FL-BPHL-1635/2020\|EPI_ISL_594324\|2020- | ···················· | ···················· | ···················· | ···················· | ···················· | ···················· |
| M | 1458 hCoV-19/USA/FL-BPHL-1643/2020\|EPI_ISL_594332\|2020- | ···················· | ···················· | ···················· | ···················· | ···················· | ···················· |
| M | 1459 hCoV-19/USA/FL-BPHL-1645/2020\|EPI_ISL_594334\|2020- | ···················· | ···················· | ···················· | ···················· | ···················· | ···················· |
| M | 1461 hCoV-19/USA/FL-BPHL-1648/2020\|EPI_ISL_594337\|2020- | ···················· | ···················· | ···················· | ···················· | ···················· | ···················· |
| M | 1462 hCoV-19/USA/FL-BPHL-1651/2020\|EPI_ISL_594340\|2020- | ···················· | ···················· | ···················· | ···················· | ···················· | ···················· |
| M | 1463 hCoV-19/USA/FL-BPHL-1652/2020\|EPI_ISL_594341\|2020- | ···················· | ···················· | ···················· | ···················· | ···················· | ···················· |
| M | 1464 hCoV-19/USA/FL-BPHL-1653/2020\|EPI_ISL_594342\|2020- | ···················· | ···················· | ···················· | ···················· | ···················· | ···················· |
| M | 1465 hCoV-19/USA/FL-BPHL-1654/2020\|EPI_ISL_594343\|2020- | ···················· | ···················· | ···················· | ···················· | ···················· | ···················· |
| M | 1466 hCoV-19/USA/FL-BPHL-1655/2020\|EPI_ISL_594344\|2020- | ··············G····· | ···················· | ···················· | ···················· | ···················· | ···················· |
| M | 1468 hCoV-19/USA/FL-BPHL-1657/2020\|EPI_ISL_594346\|2020- | ···················· | ···················· | ···················· | ···················· | ···················· | ···················· |
| M | 1469 hCoV-19/USA/FL-BPHL-1658/2020\|EPI_ISL_594347\|2020- | ···················· | ·········A·········· | ···················· | ···················· | ···················· | ···················· |
| M | 1470 hCoV-19/USA/FL-BPHL-1660/2020\|EPI_ISL_594349\|2020- | ···················· | ···················· | ···················· | ···················· | ···················· | ···················· |
| M | 1471 hCoV-19/USA/FL-BPHL-1661/2020\|EPI_ISL_594350\|2020- | ···················· | ···················· | ···················· | ···················· | ···················· | ···················· |
| M | 1472 hCoV-19/USA/FL-BPHL-1662/2020\|EPI_ISL_594351\|2020- | ···················· | ···················· | ···················· | ···················· | ···················· | ···················· |
| M | 1474 hCoV-19/USA/FL-BPHL-1664/2020\|EPI_ISL_594353\|2020- | ···················· | ···················· | ···················· | ···················· | ···················· | ···················· |
| M | 1475 hCoV-19/USA/FL-BPHL-1665/2020\|EPI_ISL_594354\|2020- | ···················· | ···················· | ···················· | ···················· | ···················· | ···················· |
| M | 1476 hCoV-19/USA/FL-BPHL-1666/2020\|EPI_ISL_594355\|2020- | ···················· | ···················· | ···················· | ···················· | ···················· | ···················· |
| M | 1477 hCoV-19/USA/FL-BPHL-1668/2020\|EPI_ISL_594357\|2020- | ···················· | ···················· | ···················· | ···················· | ···················· | ···················· |
| M | 1478 hCoV-19/USA/FL-BPHL-1671/2020\|EPI_ISL_594360\|2020- | ···················· | ···················· | ···················· | ···················· | ···················· | ···················· |
| M | 1479 hCoV-19/USA/FL-BPHL-1672/2020\|EPI_ISL_594361\|2020- | ···················· | ···················· | ···················· | ···················· | ···················· | ···················· |
| M | 1480 hCoV-19/USA/FL-BPHL-1673/2020\|EPI_ISL_594362\|2020- | ···················· | ···················· | ···················· | ···················· | ···················· | ···················· |
| M | 1481 hCoV-19/USA/FL-BPHL-1674/2020\|EPI_ISL_594363\|2020- | ···················· | ···················· | ···················· | ···················· | ···················· | ···················· |
| M | 1483 hCoV-19/USA/FL-BPHL-1680/2020\|EPI_ISL_594369\|2020- | ···················· | ···················· | ···················· | ···················· | ···················· | ···················· |
| M | 1485 hCoV-19/USA/FL-BPHL-1684/2020\|EPI_ISL_594373\|2020- | ···················· | ···················· | ···················· | ···················· | ···················· | ···················· |
| M | 1486 hCoV-19/USA/FL-BPHL-1685/2020\|EPI_ISL_594374\|2020- | ···················· | ···················· | ···················· | ···················· | ···················· | ···················· |
| M | 1487 hCoV-19/USA/FL-BPHL-1686/2020\|EPI_ISL_594375\|2020- | ···················· | ···················· | ···················· | ···················· | ···················· | ···················· |
| M | 1488 hCoV-19/USA/FL-BPHL-1699/2020\|EPI_ISL_594388\|2020- | ···················· | ···················· | ···················· | ···················· | ···················· | ···················· |
| M | 1489 hCoV-19/USA/FL-BPHL-1700/2020\|EPI_ISL_594389\|2020- | ···················· | ···················· | ···················· | ···················· | ···················· | ···················· |
| M | 1490 hCoV-19/USA/FL-BPHL-1702/2020\|EPI_ISL_594391\|2020- | ···················· | ···················· | ···················· | ···················· | ···················· | ···················· |
| M | 1491 hCoV-19/USA/UT-UPHL-201007307/2020\|EPI_ISL_594394\| | ···················· | ···················· | ···················· | ···················· | ···················· | ···················· |
| M | 1492 hCoV-19/USA/UT-UPHL-201021712/2020\|EPI_ISL_594395\| | ···················· | ···················· | ···················· | ···················· | ···················· | ···················· |
| M | 1493 hCoV-19/USA/UT-UPHL-201019448/2020\|EPI_ISL_594396\| | ···················· | ···················· | ···················· | ···················· | ···················· | ···················· |
| M | 1494 hCoV-19/USA/UT-UPHL-201016783/2020\|EPI_ISL_594398\| | ···················· | ···················· | ···················· | ···················· | ···················· | ···················· |
| M | 1495 hCoV-19/USA/UT-UPHL-201030130/2020\|EPI_ISL_594399\| | ···················· | ···················· | ···················· | ···················· | ···················· | ···················· |
| M | 1496 hCoV-19/Scotland/QEUH-9F6FE9/2020\|EPI_ISL_595652\|2 | ···················· | ···················· | ···················· | ···················· | ···················· | ···················· |
| M | 1497 hCoV-19/Scotland/QEUH-9F6FF8/2020\|EPI_ISL_595653\|2 | ···················· | ···················· | ···················· | ···················· | ···················· | ···················· |
| M | 1498 hCoV-19/Scotland/QEUH-9F7003/2020\|EPI_ISL_595654\|2 | ···················· | ···················· | ···················· | ···················· | ···················· | ···················· |
| M | 1499 hCoV-19/Scotland/QEUH-9F7021/2020\|EPI_ISL_595655\|2 | ···················· | ···················· | ···················· | ···················· | ···················· | ···················· |
| M | 1500 hCoV-19/Scotland/QEUH-9F705E/2020\|EPI_ISL_595656\|2 | ···················· | ···················· | ···················· | ···················· | ···················· | ···················· |
| M | 1501 hCoV-19/Scotland/QEUH-9F707C/2020\|EPI_ISL_595657\|2 | ···················· | ···················· | ···················· | ···················· | ···················· | ···················· |
| M | 1502 hCoV-19/England/QEUH-9F70A9/2020\|EPI_ISL_595658\|20 | ···················· | ···················· | ···················· | ···················· | ···················· | ···················· |
| M | 1503 hCoV-19/Scotland/QEUH-9F70B8/2020\|EPI_ISL_595659\|2 | ···················· | ···················· | ···················· | ···················· | ···················· | ···················· |
| M | 1504 hCoV-19/Scotland/QEUH-9F70C7/2020\|EPI_ISL_595660\|2 | ···················· | ···················· | ···················· | ···················· | ···················· | ···················· |
| M | 1505 hCoV-19/Scotland/QEUH-9F7100/2020\|EPI_ISL_595662\|2 | ···················· | ···················· | ···················· | ···················· | ···················· | ···················· |
| M | 1506 hCoV-19/Scotland/QEUH-9F711F/2020\|EPI_ISL_595663\|2 | ···················· | ···················· | ···················· | ···················· | ···················· | ···················· |
| M | 1507 hCoV-19/England/QEUH-9F712E/2020\|EPI_ISL_595664\|20 | ···················· | ···················· | ···················· | ···················· | ···················· | ···················· |

| | | | | | | | | |
|---|---|---|---|---|---|---|---|---|
| M | 1508 | hCoV-19/England/QEUH-9F713D/2020\|EPI_ISL_595665\|20 | ···················· | ···················· | ····················· | ···················· | ···················· | ···················· |
| M | 1509 | hCoV-19/England/QEUH-9F715B/2020\|EPI_ISL_595666\|20 | ···················· | ···················· | ····················· | ···················· | ···················· | ···················· |
| M | 1510 | hCoV-19/England/QEUH-9F71A6/2020\|EPI_ISL_595667\|20 | ···················· | ···················· | ····················· | ···················· | ···················· | ···················· |
| M | 1511 | hCoV-19/England/QEUH-9F71B5/2020\|EPI_ISL_595668\|20 | ···················· | ···················· | ····················· | ···················· | ···················· | ···················· |
| M | 1512 | hCoV-19/England/QEUH-9F71C4/2020\|EPI_ISL_595669\|20 | ···················· | ···················· | ····················· | ···················· | ···················· | ···················· |
| M | 1513 | hCoV-19/England/QEUH-9F71F1/2020\|EPI_ISL_595670\|20 | ···················· | ···················· | ····················· | ···················· | ···················· | ···················· |
| M | 1514 | hCoV-19/England/QEUH-9F7267/2020\|EPI_ISL_595672\|20 | ···················· | ·A·················· | ····················· | ···················· | ···················· | ···················· |
| M | 1515 | hCoV-19/England/QEUH-9F7276/2020\|EPI_ISL_595673\|20 | ···················· | ···················· | ····················· | ···················· | ···················· | ···················· |
| M | 1516 | hCoV-19/England/QEUH-9F7285/2020\|EPI_ISL_595674\|20 | ···················· | ···················· | ····················· | ···················· | ···················· | ···················· |
| M | 1517 | hCoV-19/England/QEUH-9F7294/2020\|EPI_ISL_595675\|20 | ···················· | ···················· | ····················· | ···················· | ···················· | ···················· |
| M | 1518 | hCoV-19/England/QEUH-9F72A3/2020\|EPI_ISL_595676\|20 | ···················· | ···················· | ····················· | ···················· | ···················· | ···················· |
| M | 1519 | hCoV-19/Wales/QEUH-9F72B2/2020\|EPI_ISL_595677\|2020 | ···················· | ···················· | ····················· | ···················· | ···················· | ···················· |
| M | 1520 | hCoV-19/England/QEUH-9F72EF/2020\|EPI_ISL_595678\|20 | ···················· | ···················· | ····················· | ···················· | ···················· | ···················· |
| M | 1521 | hCoV-19/Wales/QEUH-9F7319/2020\|EPI_ISL_595679\|2020 | ···················· | ···················· | ····················· | ···················· | ···················· | ···················· |
| M | 1522 | hCoV-19/England/QEUH-9F7355/2020\|EPI_ISL_595680\|20 | ···················· | ···················· | ····················· | ···················· | ···················· | ···················· |
| M | 1523 | hCoV-19/England/QEUH-9F7373/2020\|EPI_ISL_595681\|20 | ···················· | ···················· | ····················· | ···················· | ···················· | ···················· |
| M | 1524 | hCoV-19/England/QEUH-9F7391/2020\|EPI_ISL_595682\|20 | ···················· | ···················· | ····················· | ···················· | ···················· | ···················· |
| M | 1525 | hCoV-19/England/QEUH-9F73A0/2020\|EPI_ISL_595683\|20 | ···················· | ···················· | ····················· | ···················· | ····T··············· | ···················· |
| M | 1526 | hCoV-19/England/QEUH-9F73BF/2020\|EPI_ISL_595684\|20 | ···················· | ···················· | ····················· | ···················· | ····T··············· | ···················· |
| M | 1527 | hCoV-19/England/QEUH-9F73FB/2020\|EPI_ISL_595685\|20 | ···················· | ···················· | ····················· | ···················· | ···················· | ···················· |
| M | 1528 | hCoV-19/Scotland/QEUH-9F756E/2020\|EPI_ISL_595686\|2 | ···················· | ···················· | ····················· | ···················· | ···················· | ···················· |
| M | 1529 | hCoV-19/England/QEUH-9F758C/2020\|EPI_ISL_595687\|20 | ···················· | ···················· | ····················· | ···················· | ···················· | ···················· |
| M | 1530 | hCoV-19/England/QEUH-9F75AA/2020\|EPI_ISL_595688\|20 | ···················· | ···················· | ····················· | ···················· | ···················· | ···················· |
| M | 1531 | hCoV-19/England/QEUH-9F75B9/2020\|EPI_ISL_595689\|20 | ···················· | ···················· | ····················· | ···················· | ···················· | ···················· |
| M | 1532 | hCoV-19/England/QEUH-9F764D/2020\|EPI_ISL_595691\|20 | ···················· | ···················· | ····················· | ···················· | ···················· | ···················· |
| M | 1533 | hCoV-19/England/QEUH-9F765C/2020\|EPI_ISL_595692\|20 | ···················· | ···················· | ····················· | ···················· | ···················· | ···················· |
| M | 1534 | hCoV-19/England/QEUH-9F766B/2020\|EPI_ISL_595693\|20 | ···················· | ···················· | ····················· | ···················· | ···················· | ···················· |
| M | 1535 | hCoV-19/England/QEUH-9F76D4/2020\|EPI_ISL_595694\|20 | ···················· | ···················· | ····················· | ···················· | ···················· | ···················· |
| M | 1536 | hCoV-19/England/QEUH-9F76E3/2020\|EPI_ISL_595695\|20 | ···················· | ···················· | ····················· | ···················· | ····T··············· | ···················· |
| M | 1537 | hCoV-19/Brazil/SP-370/2020\|EPI_ISL_583500\|2020-06- | ···················· | ···················· | ····················· | ···················· | ···················· | ···················· |
| M | 1538 | hCoV-19/Brazil/SP-393/2020\|EPI_ISL_583502\|2020-06- | ···················· | ···················· | ····················· | ···················· | ···················· | ···················· |
| M | 1539 | hCoV-19/Brazil/SP-394/2020\|EPI_ISL_583503\|2020-06- | ···················· | ···················· | ····················· | ···················· | ···················· | ···················· |
| M | 1540 | hCoV-19/Brazil/SP-395/2020\|EPI_ISL_583504\|2020-06- | ···················· | ···················· | ····················· | ···················· | ···················· | ···················· |
| M | 1541 | hCoV-19/Brazil/SP-397/2020\|EPI_ISL_583505\|2020-06- | ···················· | ···················· | ····················· | ···················· | ···················· | ···················· |
| M | 1542 | hCoV-19/USA/MI-MDHHS-SC22049/2020\|EPI_ISL_583506\|2 | ···················· | ·A·················· | ····················· | ···················· | ···················· | ···················· |
| M | 1543 | hCoV-19/USA/MI-MDHHS-SC22050/2020\|EPI_ISL_583507\|2 | ···················· | ·············A······ | ····················· | ···················· | ···················· | ···················· |
| M | 1544 | hCoV-19/USA/MI-MDHHS-SC22051/2020\|EPI_ISL_583508\|2 | ···················· | ···················· | ····················· | ···················· | ···················· | ···················· |
| M | 1545 | hCoV-19/USA/MI-MDHHS-SC22054/2020\|EPI_ISL_583511\|2 | ···················· | ·A·················· | ····················· | ···················· | ···················· | ···················· |
| M | 1546 | hCoV-19/USA/MI-MDHHS-SC22058/2020\|EPI_ISL_583515\|2 | ···················· | ···················· | ····················· | ···················· | ···················· | ···················· |
| M | 1547 | hCoV-19/USA/MI-MDHHS-SC22060/2020\|EPI_ISL_583517\|2 | ···················· | ·A·················· | ····················· | ···················· | ···················· | ···················· |
| M | 1548 | hCoV-19/USA/MI-MDHHS-SC22061/2020\|EPI_ISL_583518\|2 | ···················· | ···················· | ····················· | ···················· | ···················· | ···················· |
| M | 1549 | hCoV-19/USA/MI-MDHHS-SC22062/2020\|EPI_ISL_583519\|2 | ···················· | ·A·················· | ····················· | ···················· | ···················· | ···················· |
| M | 1550 | hCoV-19/USA/MI-MDHHS-SC22064/2020\|EPI_ISL_583521\|2 | ···················· | ·A·················· | ····················· | ···················· | ···················· | ···················· |
| M | 1551 | hCoV-19/USA/MI-MDHHS-SC22065/2020\|EPI_ISL_583522\|2 | ···················· | ·A·················· | ····················· | ···················· | ···················· | ···················· |
| M | 1552 | hCoV-19/USA/MI-MDHHS-SC22066/2020\|EPI_ISL_583523\|2 | ···················· | ···················· | ····················· | ···················· | ···················· | ···················· |
| M | 1553 | hCoV-19/USA/MI-MDHHS-SC22068/2020\|EPI_ISL_583525\|2 | ···················· | ···················· | ····················· | ···················· | ···················· | ···················· |
| M | 1554 | hCoV-19/USA/MI-MDHHS-SC22073/2020\|EPI_ISL_583530\|2 | ···················· | ·A·················· | ····················· | ···················· | ···················· | ···················· |
| M | 1555 | hCoV-19/USA/MI-MDHHS-SC22074/2020\|EPI_ISL_583531\|2 | ···················· | ·A····A············· | ····················· | ···················· | ···················· | ···················· |
| M | 1556 | hCoV-19/USA/MI-MDHHS-SC22075/2020\|EPI_ISL_583532\|2 | ···················· | ···················· | ····················· | ···················· | ···················· | ···················· |
| M | 1557 | hCoV-19/England/NOTT-115201/2020\|EPI_ISL_584801\|20 | ···················· | ···················· | ····················· | ···················· | ···················· | ···················· |
| M | 1558 | hCoV-19/England/NOTT-11531D/2020\|EPI_ISL_584802\|20 | ···················· | ···················· | ····················· | ···················· | ···················· | ···················· |
| M | 1559 | hCoV-19/England/NOTT-11533B/2020\|EPI_ISL_584804\|20 | ···················· | ···················· | ····················· | ···················· | ···················· | ···················· |
| M | 1560 | hCoV-19/England/NOTT-115359/2020\|EPI_ISL_584805\|20 | ···················· | ···················· | ····················· | ···················· | ···················· | ···················· |
| M | 1561 | hCoV-19/England/NOTT-115368/2020\|EPI_ISL_584806\|20 | ···················· | ···················· | ····················· | ···················· | ···················· | ···················· |
| M | 1562 | hCoV-19/England/NOTT-115386/2020\|EPI_ISL_584808\|20 | ···················· | ···················· | ····················· | ···················· | ···················· | ···················· |
| M | 1563 | hCoV-19/England/QEUH-9DE925/2020\|EPI_ISL_584812\|20 | ···················· | ···················· | ····················· | ···················· | ···················· | ···················· |
| M | 1564 | hCoV-19/England/QEUH-9DE934/2020\|EPI_ISL_584813\|20 | ···················· | ···················· | ····················· | ···················· | ···················· | ···················· |
| M | 1565 | hCoV-19/England/QEUH-9DE970/2020\|EPI_ISL_584815\|20 | ···················· | ···················· | ····················· | ···················· | ···················· | ···················· |
| M | 1566 | hCoV-19/England/QEUH-9DEB2F/2020\|EPI_ISL_584818\|20 | ···················· | ···················· | ····················· | ···················· | ···················· | ···················· |
| M | 1567 | hCoV-19/England/QEUH-9DEB5C/2020\|EPI_ISL_584819\|20 | ···················· | ···················· | ····················· | ···················· | ···················· | ···················· |
| M | 1568 | hCoV-19/England/QEUH-9DEB6B/2020\|EPI_ISL_584820\|20 | ···················· | ···················· | ····················· | ···················· | ······T············· | ···················· |
| M | 1569 | hCoV-19/England/QEUH-9DEC3B/2020\|EPI_ISL_584821\|20 | ···················· | ···················· | ····················· | ···················· | ···················· | ···················· |
| M | 1570 | hCoV-19/England/QEUH-9DEC95/2020\|EPI_ISL_584822\|20 | ···················· | ·A·················· | ····················· | ···················· | ···················· | ···A················ |
| M | 1571 | hCoV-19/England/QEUH-9DECB3/2020\|EPI_ISL_584823\|20 | ···················· | ···················· | ····················· | ···················· | ······T············· | ···················· |
| M | 1572 | hCoV-19/England/QEUH-9DECFF/2020\|EPI_ISL_584824\|20 | ···················· | ···················· | ····················· | ···················· | ···················· | ···················· |
| M | 1573 | hCoV-19/England/QEUH-9DED1A/2020\|EPI_ISL_584825\|20 | ···················· | ···················· | ····················· | ···················· | ···················· | ···················· |
| M | 1574 | hCoV-19/England/QEUH-9DED29/2020\|EPI_ISL_584826\|20 | ···················· | ···················· | ····················· | ···················· | ···················· | ···················· |
| M | 1575 | hCoV-19/England/QEUH-9DED83/2020\|EPI_ISL_584827\|20 | ···················· | ···················· | ····················· | ···················· | ···················· | ···················· |
| M | 1576 | hCoV-19/England/QEUH-9DEE26/2020\|EPI_ISL_584828\|20 | ···················· | ···················· | ····················· | ···················· | ···················· | ···················· |
| M | 1577 | hCoV-19/England/QEUH-9DEE53/2020\|EPI_ISL_584830\|20 | ···················· | ·A·················· | ····················· | ···················· | ···················· | ···A················ |
| M | 1578 | hCoV-19/England/QEUH-9DEE62/2020\|EPI_ISL_584831\|20 | ···················· | ···················· | ····················· | ···················· | ···················· | ···················· |
| M | 1579 | hCoV-19/England/QEUH-9DEE71/2020\|EPI_ISL_584832\|20 | ···················· | ···················· | ····················· | ···················· | ···················· | ···················· |
| M | 1580 | hCoV-19/England/QEUH-9DEF32/2020\|EPI_ISL_584833\|20 | ···················· | ···················· | ····················· | ···················· | ···················· | ···················· |
| M | 1581 | hCoV-19/England/QEUH-9DEF7E/2020\|EPI_ISL_584835\|20 | ···················· | ···················· | ····················· | ···················· | ···················· | ···················· |
| M | 1582 | hCoV-19/England/QEUH-9DEF9C/2020\|EPI_ISL_584836\|20 | ···················· | ···················· | ····················· | ···················· | ···················· | ···················· |
| M | 1583 | hCoV-19/England/QEUH-9DEFBA/2020\|EPI_ISL_584837\|20 | ···················· | ···················· | ····················· | ···················· | ···················· | ···················· |
| M | 1584 | hCoV-19/England/QEUH-9DEFD8/2020\|EPI_ISL_584839\|20 | ···················· | ···················· | ····················· | ···················· | ···················· | ···················· |
| M | 1585 | hCoV-19/England/QEUH-9DEFE7/2020\|EPI_ISL_584840\|20 | ···················· | ···················· | ····················· | ···················· | ······T············· | ···················· |
| M | 1586 | hCoV-19/England/QEUH-9DF001/2020\|EPI_ISL_584841\|20 | ···················· | ···················· | ····················· | ···················· | ···················· | ···················· |
| M | 1587 | hCoV-19/England/QEUH-9DF010/2020\|EPI_ISL_584842\|20 | ···················· | ···················· | ····················· | ···················· | ····T··············· | ···················· |

| | | | | | | | |
|---|---|---|---|---|---|---|---|
| M | 1588 hCoV-19/England/QEUH-9DF03E/2020\|EPI_ISL_584843\|20 | .................... | .................... | .................... | .................... | .................... | .................... |
| M | 1589 hCoV-19/England/QEUH-9DF0D4/2020\|EPI_ISL_584846\|20 | .................... | .................... | .................... | .................... | .................... | .................... |
| M | 1590 hCoV-19/England/QEUH-9E3495/2020\|EPI_ISL_584847\|20 | .................... | .A.................. | .................... | .................... | .................... | ..A................. |
| M | 1591 hCoV-19/England/QEUH-9E34A4/2020\|EPI_ISL_584848\|20 | .................... | .................... | .................... | .................... | .................... | .................... |
| M | 1592 hCoV-19/England/QEUH-9E35FC/2020\|EPI_ISL_584858\|20 | .................... | .................... | .................... | .................... | .................... | .................... |
| M | 1593 hCoV-19/England/QEUH-9E3626/2020\|EPI_ISL_584859\|20 | .................... | .................... | .................... | .................... | .................... | .................... |
| M | 1594 hCoV-19/England/QEUH-9E3653/2020\|EPI_ISL_584860\|20 | .................... | .................... | .................... | .................... | .................... | .................... |
| M | 1595 hCoV-19/England/QEUH-9E3662/2020\|EPI_ISL_584861\|20 | .................... | .................... | .................... | .................... | .....T.............. | .................... |
| M | 1596 hCoV-19/England/QEUH-9E3671/2020\|EPI_ISL_584862\|20 | .................... | .A.................. | .................... | .................... | .................... | .................... |
| M | 1597 hCoV-19/England/QEUH-9E36DB/2020\|EPI_ISL_584865\|20 | .................... | .................... | .................... | .................... | .................... | .................... |
| M | 1598 hCoV-19/England/QEUH-9E36EA/2020\|EPI_ISL_584866\|20 | .................... | .................... | .................... | .................... | .................... | .................... |
| M | 1599 hCoV-19/England/QEUH-9E3750/2020\|EPI_ISL_584869\|20 | .................... | .................... | .................... | .................... | .................... | .................... |
| M | 1600 hCoV-19/England/QEUH-9E378D/2020\|EPI_ISL_584870\|20 | .................... | .................... | .................... | .................... | .................... | .................... |
| M | 1601 hCoV-19/England/QEUH-9E379C/2020\|EPI_ISL_584871\|20 | .................... | .................... | .................... | .................... | .................... | .................... |
| M | 1602 hCoV-19/England/QEUH-9E37AB/2020\|EPI_ISL_584872\|20 | .................... | .................... | .................... | .................... | .................... | .................... |
| M | 1603 hCoV-19/England/QEUH-9E37BA/2020\|EPI_ISL_584873\|20 | .................... | .................... | .................... | .................... | .................... | .................... |
| M | 1604 hCoV-19/England/QEUH-9E37F6/2020\|EPI_ISL_584875\|20 | .................... | .A.................. | .................... | .................... | .................... | .................... |
| M | 1605 hCoV-19/England/QEUH-9E387B/2020\|EPI_ISL_584876\|20 | .................... | .................... | .................... | .................... | .................... | .................... |
| M | 1606 hCoV-19/England/QEUH-9E394B/2020\|EPI_ISL_584877\|20 | .................... | .................... | .................... | .................... | .................... | .................... |
| M | 1607 hCoV-19/England/QEUH-9E3969/2020\|EPI_ISL_584879\|20 | .................... | .A.................. | .................... | .................... | .................... | .................... |
| M | 1608 hCoV-19/England/QEUH-9E3978/2020\|EPI_ISL_584880\|20 | .................... | .................... | .................... | .................... | .................... | .................... |
| M | 1609 hCoV-19/England/QEUH-9E39C3/2020\|EPI_ISL_584881\|20 | .................... | .................... | .................... | .................... | .................... | .................... |
| M | 1610 hCoV-19/England/ALDP-A2EA7D/2020\|EPI_ISL_587000\|20 | .................... | .................... | .................... | .................... | .................... | .................... |
| M | 1611 hCoV-19/England/ALDP-A2EAF5/2020\|EPI_ISL_587001\|20 | .................... | .................... | .................... | .................... | .................... | .................... |
| M | 1612 hCoV-19/England/ALDP-A2F6B4/2020\|EPI_ISL_587004\|20 | .................... | .................... | .................... | .................... | .................... | .................... |
| M | 1613 hCoV-19/England/ALDP-A2FCD0/2020\|EPI_ISL_587005\|20 | .................... | .................... | .................... | .................... | .................... | .................... |
| M | 1614 hCoV-19/England/ALDP-A2EB01/2020\|EPI_ISL_587017\|20 | .................... | .................... | .................... | .................... | .................... | .................... |
| M | 1615 hCoV-19/England/ALDP-A2F1C2/2020\|EPI_ISL_587018\|20 | .................... | .................... | .................... | .................... | .................... | .................... |
| M | 1616 hCoV-19/England/ALDP-A2F0B6/2020\|EPI_ISL_587019\|20 | .................... | .................... | .................... | .................... | .................... | .................... |
| M | 1617 hCoV-19/England/ALDP-A2F8FA/2020\|EPI_ISL_587020\|20 | .................... | .................... | .................... | .................... | .................... | .................... |
| M | 1618 hCoV-19/England/ALDP-A2F326/2020\|EPI_ISL_587021\|20 | .................... | .................... | .................... | .................... | .................... | .................... |
| M | 1619 hCoV-19/England/ALDP-A2F5A8/2020\|EPI_ISL_587022\|20 | .................... | .................... | .................... | .................... | .................... | .................... |
| M | 1620 hCoV-19/England/ALDP-A2FB5B/2020\|EPI_ISL_587023\|20 | .................... | .................... | .................... | .................... | .................... | .................... |
| M | 1621 hCoV-19/England/ALDP-A2FD0A/2020\|EPI_ISL_587024\|20 | .................... | .................... | .................... | .................... | .................... | .................... |
| M | 1622 hCoV-19/England/ALDP-A2EDA1/2020\|EPI_ISL_587025\|20 | .................... | .................... | .................... | .................... | .................... | .................... |
| M | 1623 hCoV-19/England/ALDP-A2F7FD/2020\|EPI_ISL_587026\|20 | .................... | .................... | .................... | .................... | .................... | .................... |
| M | 1624 hCoV-19/England/ALDP-A2ED92/2020\|EPI_ISL_587027\|20 | .................... | .................... | .................... | .................... | .................... | .................... |
| M | 1625 hCoV-19/England/ALDP-A2EA40/2020\|EPI_ISL_587028\|20 | .................... | .................... | .................... | .................... | .................... | .................... |
| M | 1626 hCoV-19/England/ALDP-A2FEE9/2020\|EPI_ISL_587029\|20 | .................... | .................... | .................... | .................... | .................... | .................... |
| M | 1627 hCoV-19/England/ALDP-A2F371/2020\|EPI_ISL_587030\|20 | .................... | .................... | .................... | .................... | .................... | .................... |
| M | 1628 hCoV-19/England/ALDP-A2FEBC/2020\|EPI_ISL_587031\|20 | .................... | .................... | .................... | .................... | .................... | .................... |
| M | 1629 hCoV-19/England/ALDP-A2F951/2020\|EPI_ISL_587034\|20 | .................... | .................... | .................... | .................... | .................... | .................... |
| M | 1630 hCoV-19/England/ALDP-A2FA03/2020\|EPI_ISL_587035\|20 | .................... | .................... | .................... | .................... | .................... | .................... |
| M | 1631 hCoV-19/England/ALDP-A2FDCE/2020\|EPI_ISL_587036\|20 | .................... | .................... | .................... | .................... | .................... | .................... |
| M | 1632 hCoV-19/England/ALDP-A2F432/2020\|EPI_ISL_587037\|20 | .................... | .................... | .................... | .................... | .................... | .................... |
| M | 1633 hCoV-19/England/ALDP-A2FC76/2020\|EPI_ISL_587038\|20 | .................... | .................... | .................... | .................... | .................... | .................... |
| M | 1634 hCoV-19/England/ALDP-A2F2CF/2020\|EPI_ISL_587080\|20 | .................... | .................... | .................... | .................... | .................... | .................... |
| M | 1635 hCoV-19/England/ALDP-A2F2FC/2020\|EPI_ISL_587081\|20 | .................... | .................... | .................... | .................... | .................... | .................... |
| M | 1636 hCoV-19/England/ALDP-A2FF04/2020\|EPI_ISL_587082\|20 | .................... | .................... | .................... | .................... | .................... | .................... |
| M | 1637 hCoV-19/England/ALDP-A2E99E/2020\|EPI_ISL_587083\|20 | .................... | .A.................. | .................... | .................... | .................... | ..A................. |
| M | 1638 hCoV-19/England/ALDP-A2FF6E/2020\|EPI_ISL_587084\|20 | .................... | .................... | .................... | .................... | .................... | .................... |
| M | 1639 hCoV-19/England/ALDP-A2F5C6/2020\|EPI_ISL_587086\|20 | .................... | .................... | .................... | .................... | .................... | .................... |
| M | 1640 hCoV-19/England/ALDP-A2EB10/2020\|EPI_ISL_587089\|20 | .................... | .................... | .................... | .................... | .................... | .................... |
| M | 1641 hCoV-19/England/ALDP-A2EF50/2020\|EPI_ISL_587090\|20 | .................... | .................... | .................... | .................... | .....T.............. | .................... |
| M | 1642 hCoV-19/England/ALDP-A2F696/2020\|EPI_ISL_587091\|20 | .................... | .................... | .................... | .................... | .................... | .................... |
| M | 1643 hCoV-19/England/ALDP-A2E891/2020\|EPI_ISL_587092\|20 | .................... | .................... | .................... | .................... | .................... | .................... |
| M | 1644 hCoV-19/England/ALDP-A2FDDD/2020\|EPI_ISL_587093\|20 | .................... | .................... | .................... | .................... | .................... | .................... |
| M | 1645 hCoV-19/England/ALDP-A2F5F3/2020\|EPI_ISL_587094\|20 | .................... | .................... | .................... | .................... | .................... | .................... |
| M | 1646 hCoV-19/England/ALDP-A2ED29/2020\|EPI_ISL_587095\|20 | .................... | .................... | .................... | .................... | .................... | .................... |
| M | 1647 hCoV-19/England/ALDP-A2EC0E/2020\|EPI_ISL_587096\|20 | .................... | .................... | .................... | .................... | .................... | .................... |
| M | 1648 hCoV-19/England/ALDP-A2E9F8/2020\|EPI_ISL_587097\|20 | .................... | .................... | .................... | .................... | .................... | .................... |
| M | 1649 hCoV-19/England/ALDP-A2E916/2020\|EPI_ISL_587098\|20 | .................... | .A.................. | .................... | .................... | .................... | .................... |
| M | 1650 hCoV-19/England/ALDP-A2FA6D/2020\|EPI_ISL_587099\|20 | .................... | .................... | .................... | .................... | .................... | .................... |
| M | 1651 hCoV-19/England/QEUH-9F5A9E/2020\|EPI_ISL_588326\|20 | .................... | .................... | .................... | .................... | .................... | .................... |
| M | 1652 hCoV-19/England/QEUH-9F48FF/2020\|EPI_ISL_588327\|20 | ........T........... | .................... | .................... | .................... | .................... | .................... |
| M | 1653 hCoV-19/England/QEUH-9F5D0E/2020\|EPI_ISL_588328\|20 | .................... | .................... | .................... | .................... | .................... | .................... |
| M | 1654 hCoV-19/England/QEUH-9F590A/2020\|EPI_ISL_588329\|20 | .................... | .................... | .................... | .................... | .................... | .................... |
| M | 1655 hCoV-19/England/QEUH-9F5CE3/2020\|EPI_ISL_588330\|20 | .................... | .A.................. | .................... | .................... | .................... | .................... |
| M | 1656 hCoV-19/England/QEUH-9F523C/2020\|EPI_ISL_588331\|20 | .................... | .................... | .................... | .................... | .................... | .................... |
| M | 1657 hCoV-19/England/QEUH-9F5ABC/2020\|EPI_ISL_588332\|20 | .................... | .................... | .................... | .................... | .................... | .................... |
| M | 1658 hCoV-19/England/QEUH-9F5D4A/2020\|EPI_ISL_588335\|20 | .................... | .................... | .................... | .................... | .................... | .................... |
| M | 1659 hCoV-19/England/QEUH-9F580D/2020\|EPI_ISL_588336\|20 | .................... | .................... | .................... | .................... | .................... | .................... |
| M | 1660 hCoV-19/England/QEUH-9F5C2F/2020\|EPI_ISL_588348\|20 | .................... | .A.......A.......... | .................... | .................... | .................... | .................... |
| M | 1661 hCoV-19/England/QEUH-9F5C7A/2020\|EPI_ISL_588350\|20 | .................... | .................... | .................... | .................... | .................... | .................... |
| M | 1662 hCoV-19/England/QEUH-9F6909/2020\|EPI_ISL_588351\|20 | .................... | .................... | .................... | .................... | .................... | .................... |
| M | 1663 hCoV-19/England/QEUH-9F47B6/2020\|EPI_ISL_588352\|20 | .................... | .................... | .................... | .................... | .................... | .................... |
| M | 1664 hCoV-19/England/QEUH-9F51E4/2020\|EPI_ISL_588353\|20 | .................... | .................... | .................... | .................... | .................... | .................... |
| M | 1665 hCoV-19/England/QEUH-9F5CF2/2020\|EPI_ISL_588354\|20 | .................... | .................... | .................... | .................... | .................... | .................... |
| M | 1666 hCoV-19/England/QEUH-9F5946/2020\|EPI_ISL_588355\|20 | .................... | .................... | .................... | .................... | .................... | .................... |
| M | 1667 hCoV-19/England/QEUH-9F5BD7/2020\|EPI_ISL_588356\|20 | .................... | .................... | .................... | .................... | .................... | .................... |

| | | | | | | | | |
|---|---|---|---|---|---|---|---|---|
| M | 1668 hCoV-19/England/QEUH-9F4877/2020\|EPI_ISL_588357\|20 | ···················· | ···················· | ···················· | ···················· | ···················· | ···················· | |
| M | 1669 hCoV-19/England/QEUH-9F4992/2020\|EPI_ISL_588358\|20 | ···················· | ···················· | ···················· | ···················· | ···················· | ···················· | |
| M | 1670 hCoV-19/England/QEUH-9F4B23/2020\|EPI_ISL_588359\|20 | ···················· | ···················· | ···················· | ···················· | ····T··············· | ···················· | |
| M | 1671 hCoV-19/England/QEUH-9F5D3B/2020\|EPI_ISL_588361\|20 | ···················· | ···················· | ···················· | ···················· | ···················· | ···················· | |
| M | 1672 hCoV-19/England/QEUH-9F5E1A/2020\|EPI_ISL_588362\|20 | ···················· | ···················· | ···················· | ···················· | ···················· | ···················· | |
| M | 1673 hCoV-19/England/QEUH-9F64BD/2020\|EPI_ISL_588363\|20 | ···················· | ···················· | ···················· | ···················· | ···················· | ···················· | |
| M | 1674 hCoV-19/England/QEUH-9F4561/2020\|EPI_ISL_588364\|20 | ···················· | ···················· | ···················· | ···················· | ···················· | ···················· | |
| M | 1675 hCoV-19/England/QEUH-9F480E/2020\|EPI_ISL_588365\|20 | ···················· | ···················· | ···················· | ···················· | ····T··············· | ···················· | |
| M | 1676 hCoV-19/England/QEUH-9F48B3/2020\|EPI_ISL_588366\|20 | ···················· | ···················· | ···················· | ···················· | ···················· | ···················· | |
| M | 1677 hCoV-19/England/QEUH-9F484A/2020\|EPI_ISL_588367\|20 | ···················· | ···················· | ···················· | ···················· | ···················· | ···················· | |
| M | 1678 hCoV-19/England/QEUH-9F582B/2020\|EPI_ISL_588368\|20 | ···················· | ···················· | ···················· | ···················· | ···················· | ···················· | |
| M | 1679 hCoV-19/England/QEUH-9F5B8C/2020\|EPI_ISL_588369\|20 | ···················· | ···················· | ···················· | ···················· | ···················· | ···················· | |
| M | 1680 hCoV-19/England/MILK-9F1EA5/2020\|EPI_ISL_589657\|20 | ···················· | ···················· | ···················· | ···················· | ···················· | ···················· | |
| M | 1681 hCoV-19/England/MILK-9F17E6/2020\|EPI_ISL_589658\|20 | ···················· | ···················· | ···················· | ···················· | ···················· | ···················· | |
| M | 1682 hCoV-19/England/MILK-9F0DE5/2020\|EPI_ISL_589659\|20 | ···················· | ···················· | ···················· | ···················· | ···················· | ···················· | |
| M | 1683 hCoV-19/England/MILK-9F1AED/2020\|EPI_ISL_589660\|20 | ···················· | ···················· | ···················· | ···················· | ···················· | ···················· | |
| M | 1684 hCoV-19/England/MILK-9F1D20/2020\|EPI_ISL_589661\|20 | ···················· | ···················· | ···················· | ···················· | ···················· | ···················· | |
| M | 1685 hCoV-19/England/MILK-9F1E1E/2020\|EPI_ISL_589662\|20 | ···················· | ···················· | ···················· | ···················· | ···················· | ···················· | |
| M | 1686 hCoV-19/England/MILK-9F1ACF/2020\|EPI_ISL_589666\|20 | ···················· | ···················· | ···················· | ···················· | ···················· | ···················· | |
| M | 1687 hCoV-19/England/MILK-9F0C7F/2020\|EPI_ISL_589667\|20 | ···················· | ···················· | ···················· | ···················· | ···················· | ···················· | |
| M | 1688 hCoV-19/England/MILK-9F0FFE/2020\|EPI_ISL_589679\|20 | ···················· | ···················· | ···················· | ···················· | ···················· | ···················· | |
| M | 1689 hCoV-19/England/MILK-9F1B08/2020\|EPI_ISL_589680\|20 | ···················· | ···················· | ···················· | ···················· | ···················· | ···················· | |
| M | 1690 hCoV-19/Wales/MILK-9F1009/2020\|EPI_ISL_589681\|2020 | ···················· | ···················· | ···················· | ···················· | ···················· | ···················· | |
| M | 1691 hCoV-19/England/MILK-9F08C6/2020\|EPI_ISL_589684\|20 | ···················· | ···················· | ···················· | ···················· | ···················· | ···················· | |
| M | 1692 hCoV-19/England/MILK-9F18E3/2020\|EPI_ISL_589685\|20 | ···················· | ···················· | ···················· | ···················· | ······T············· | ···················· | |
| M | 1693 hCoV-19/England/MILK-9F1E4B/2020\|EPI_ISL_589687\|20 | ···················· | ···················· | ···················· | ···················· | ···················· | ···················· | |
| M | 1694 hCoV-19/England/MILK-9F1045/2020\|EPI_ISL_589688\|20 | ···················· | ···················· | ···················· | ···················· | ···················· | ···················· | |
| M | 1695 hCoV-19/England/MILK-9F1EF0/2020\|EPI_ISL_589689\|20 | ···················· | ···················· | ···················· | ···················· | ···················· | ···················· | |
| M | 1696 hCoV-19/England/MILK-9F0B09/2020\|EPI_ISL_589690\|20 | ···················· | ···················· | ···················· | ···················· | ···················· | ···················· | |
| M | 1697 hCoV-19/England/MILK-9F1B17/2020\|EPI_ISL_589691\|20 | ···················· | ·A·················· | ···················· | ···················· | ···················· | ···················· | |
| M | 1698 hCoV-19/England/MILK-9F0AC0/2020\|EPI_ISL_589692\|20 | ···················· | ···················· | ···················· | ···················· | ···················· | ···················· | |
| M | 1699 hCoV-19/England/MILK-9F1E96/2020\|EPI_ISL_589693\|20 | ···················· | ···················· | ···················· | ···················· | ···················· | ···················· | |
| M | 1700 hCoV-19/England/MILK-9F1C7E/2020\|EPI_ISL_589694\|20 | ···················· | ···················· | ···················· | ···················· | ···················· | ···················· | |
| M | 1701 hCoV-19/England/MILK-9F1E3C/2020\|EPI_ISL_589696\|20 | ···················· | ···················· | ···················· | ···················· | ···················· | ···················· | |
| M | 1702 hCoV-19/England/MILK-9F1F93/2020\|EPI_ISL_589697\|20 | ···················· | ···················· | ···················· | ···················· | ···················· | ···················· | |
| M | 1703 hCoV-19/England/MILK-9F12D6/2020\|EPI_ISL_589698\|20 | ···················· | ···················· | ···················· | ···················· | ···················· | ···················· | |
| M | 1704 hCoV-19/Wales/MILK-9F1C05/2020\|EPI_ISL_589699\|2020 | ···················· | ···················· | ···················· | ···················· | ···················· | ···················· | |
| M | 1705 hCoV-19/England/QEUH-9B6EF7/2020\|EPI_ISL_590640\|20 | ···················· | ···················· | ···················· | ···················· | ···················· | ···················· | |
| M | 1706 hCoV-19/England/QEUH-9B74C7/2020\|EPI_ISL_590641\|20 | ···················· | ···················· | ···················· | ···················· | ···················· | ···················· | |
| M | 1707 hCoV-19/England/QEUH-9B8147/2020\|EPI_ISL_590642\|20 | ···················· | ···················· | ···················· | ···················· | ···················· | ···················· | |
| M | 1708 hCoV-19/England/QEUH-9B7FE4/2020\|EPI_ISL_590643\|20 | ···················· | ···················· | ···················· | ···················· | ···················· | ···················· | |
| M | 1709 hCoV-19/England/QEUH-9B7263/2020\|EPI_ISL_590660\|20 | ···················· | ···················· | ···················· | ···················· | ···················· | ···················· | |
| M | 1710 hCoV-19/England/QEUH-9B7F3F/2020\|EPI_ISL_590661\|20 | ···················· | ···················· | ···················· | ···················· | ···················· | ···················· | |
| M | 1711 hCoV-19/England/QEUH-9B72EB/2020\|EPI_ISL_590662\|20 | ···················· | ···················· | ···················· | ···················· | ···················· | ···················· | |
| M | 1712 hCoV-19/England/QEUH-9B7157/2020\|EPI_ISL_590663\|20 | ···················· | ···················· | ···················· | ···················· | ···················· | ···················· | |
| M | 1713 hCoV-19/England/QEUH-9B77FB/2020\|EPI_ISL_590665\|20 | ···················· | ···················· | ···················· | ···················· | ····T··············· | ···················· | |
| M | 1714 hCoV-19/England/QEUH-9B80C2/2020\|EPI_ISL_590666\|20 | ···················· | ·A·················· | ···················· | ···················· | ···················· | ···················· | |
| M | 1715 hCoV-19/NorthernIreland/QEUH-9B8192/2020\|EPI_ISL_5 | ···················· | ···················· | ···················· | ···················· | ···················· | ···················· | |
| M | 1716 hCoV-19/England/QEUH-9B74D6/2020\|EPI_ISL_590668\|20 | ···················· | ···················· | ···················· | ···················· | ···················· | ···················· | |
| M | 1717 hCoV-19/England/QEUH-9B78AD/2020\|EPI_ISL_590669\|20 | ···················· | ·A·················· | ···················· | ···················· | ···················· | ···················· | |
| M | 1718 hCoV-19/England/QEUH-9B6EE8/2020\|EPI_ISL_590670\|20 | ···················· | ···················· | ···················· | ···················· | ···················· | ···················· | |
| M | 1719 hCoV-19/England/QEUH-9B6FA9/2020\|EPI_ISL_590671\|20 | ···················· | ···················· | ···················· | ···················· | ···················· | ···················· | |
| M | 1720 hCoV-19/England/QEUH-9B7E50/2020\|EPI_ISL_590672\|20 | ···················· | ···················· | ···················· | ···················· | ···················· | ···················· | |
| M | 1721 hCoV-19/England/QEUH-9B7DBD/2020\|EPI_ISL_590673\|20 | ···················· | ···················· | ···················· | ···················· | ···················· | ···················· | |
| M | 1722 hCoV-19/England/QEUH-9B7DDB/2020\|EPI_ISL_590674\|20 | ···················· | ···················· | ···················· | ···················· | ···················· | ···················· | |
| M | 1723 hCoV-19/England/QEUH-9B7306/2020\|EPI_ISL_590675\|20 | ···················· | ···················· | ···················· | ···················· | ···················· | ···················· | |
| M | 1724 hCoV-19/England/QEUH-9B7F5D/2020\|EPI_ISL_590676\|20 | ···················· | ···················· | ···················· | ···················· | ···················· | ···················· | |
| M | 1725 hCoV-19/England/QEUH-9B810B/2020\|EPI_ISL_590677\|20 | ···················· | ···················· | ···················· | ···················· | ···················· | ···················· | |
| M | 1726 hCoV-19/England/QEUH-9B7C74/2020\|EPI_ISL_590678\|20 | ···················· | ·A·······A·········· | ···················· | ···················· | ···················· | ···················· | |
| M | 1727 hCoV-19/England/QEUH-9B8086/2020\|EPI_ISL_590679\|20 | ···················· | ···················· | ···················· | ···················· | ···················· | ···················· | |
| M | 1728 hCoV-19/Australia/VIC12378/2020\|EPI_ISL_591971\|202 | ···················· | ···················· | ···················· | ···················· | ···················· | ···················· | |
| M | 1729 hCoV-19/Australia/VIC12382/2020\|EPI_ISL_591972\|202 | ···················· | ···················· | ···················· | ···················· | ···················· | ···················· | |
| M | 1730 hCoV-19/Australia/VIC12383/2020\|EPI_ISL_591973\|202 | ···················· | ···················· | ···················· | ···················· | ···················· | ···················· | |
| M | 1731 hCoV-19/Australia/VIC12387/2020\|EPI_ISL_591974\|202 | ···················· | ···················· | ···················· | ···················· | ···················· | ···················· | |
| M | 1732 hCoV-19/Australia/VIC12443/2020\|EPI_ISL_591990\|202 | ···················· | ···················· | ···················· | ···················· | ···················· | ···················· | |
| M | 1733 hCoV-19/Australia/VIC12450/2020\|EPI_ISL_591991\|202 | ···················· | ···················· | ···················· | ···················· | ···················· | ···················· | |
| M | 1734 hCoV-19/Australia/VIC12451/2020\|EPI_ISL_591992\|202 | ···················· | ···················· | ···················· | ···················· | ···················· | ···················· | |
| M | 1735 hCoV-19/Australia/VIC12452/2020\|EPI_ISL_591993\|202 | ···················· | ···················· | ···················· | ···················· | NNN················· | ···················· | 1 |
| M | 1736 hCoV-19/Australia/VIC12458/2020\|EPI_ISL_591994\|202 | ···················· | ···················· | ···················· | ···················· | ···················· | ···················· | |
| M | 1737 hCoV-19/Australia/VIC12460/2020\|EPI_ISL_591995\|202 | ···················· | ···················· | ···················· | ···················· | ···················· | ···················· | |
| M | 1739 hCoV-19/Australia/VIC12462/2020\|EPI_ISL_591997\|202 | ···················· | ···················· | ···················· | ···················· | ···················· | ···················· | |
| M | 1740 hCoV-19/Australia/VIC12466/2020\|EPI_ISL_591998\|202 | ···················· | ···················· | ···················· | ···················· | ···················· | ···················· | |
| M | 1742 hCoV-19/Australia/VIC14862/2020\|EPI_ISL_593034\|202 | ···················· | ···················· | ···················· | ···················· | ···················· | ···················· | |
| M | 1744 hCoV-19/Australia/VIC14868/2020\|EPI_ISL_593036\|202 | ···················· | ···················· | ···················· | ···················· | ···················· | ···················· | |
| M | 1745 hCoV-19/Australia/VIC14869/2020\|EPI_ISL_593037\|202 | ···················· | ···················· | ···················· | ···················· | ···················· | ···················· | |
| M | 1746 hCoV-19/Australia/VIC14872/2020\|EPI_ISL_593038\|202 | ···················· | ···················· | ···················· | ···················· | ···················· | ···················· | |
| M | 1748 hCoV-19/Australia/VIC14889/2020\|EPI_ISL_593045\|202 | ···················· | ···················· | ···················· | ···················· | ···················· | ···················· | |
| M | 1749 hCoV-19/Australia/VIC14891/2020\|EPI_ISL_593046\|202 | ···················· | ···················· | ···················· | ···················· | ···················· | ···················· | |
| M | 1750 hCoV-19/Australia/VIC14892/2020\|EPI_ISL_593047\|202 | ···················· | ···················· | ···················· | ···················· | ···················· | ···················· | |
| M | 1751 hCoV-19/Australia/VIC14893/2020\|EPI_ISL_593048\|202 | ···················· | ···················· | ···················· | ···················· | ···················· | ···················· | |

| | | | | | | | | |
|---|---|---|---|---|---|---|---|---|
| M | 1753 hCoV-19/Australia/VIC14895/2020\|EPI_ISL_593050\|202 | ···················· | ···················· | ···················· | ···················· | ···················· | ···················· | |
| M | 1757 hCoV-19/Australia/VIC14902/2020\|EPI_ISL_593054\|202 | ···················· | ···················· | ···················· | ···················· | ···················· | ···················· | |
| M | 1759 hCoV-19/USA/FL-BPHL-1676/2020\|EPI_ISL_594365\|2020- | ···················· | ···················· | ···················· | ···················· | ···················· | ···················· | |
| M | 1760 hCoV-19/USA/FL-BPHL-1677/2020\|EPI_ISL_594366\|2020- | ···················· | ···················· | ···················· | ···················· | ···················· | ···················· | |
| M | 1761 hCoV-19/USA/FL-BPHL-1678/2020\|EPI_ISL_594367\|2020- | ···················· | ···A················ | ···················· | ···················· | ···················· | ···················· | |
| M | 1762 hCoV-19/USA/FL-BPHL-1679/2020\|EPI_ISL_594368\|2020- | ···················· | ···················· | ···················· | ···················· | ···················· | ···················· | |
| M | 1763 hCoV-19/USA/FL-BPHL-1687/2020\|EPI_ISL_594376\|2020- | ···················· | ···················· | ···················· | ···················· | ···················· | ···················· | |
| M | 1764 hCoV-19/USA/FL-BPHL-1689/2020\|EPI_ISL_594378\|2020- | ···················· | ···················· | ···················· | ···················· | ···················· | ···················· | |
| M | 1765 hCoV-19/USA/FL-BPHL-1691/2020\|EPI_ISL_594380\|2020- | ···················· | ···················· | ···················· | ···················· | NNN················· | ···················· | 1 |
| M | 1767 hCoV-19/Wales/PHWC-47E79B/2020\|EPI_ISL_595500\|2020 | ···················· | ···················· | ···················· | ···················· | ···················· | ···················· | |
| M | 1768 hCoV-19/Wales/PHWC-47E7AA/2020\|EPI_ISL_595501\|2020 | ···················· | ···················· | ···················· | ···················· | ···················· | ···················· | |
| M | 1769 hCoV-19/Wales/PHWC-47E7B9/2020\|EPI_ISL_595502\|2020 | ···················· | ···················· | ···················· | ···················· | ···················· | ···················· | |
| M | 1770 hCoV-19/Wales/PHWC-47E7D7/2020\|EPI_ISL_595504\|2020 | ···················· | ···················· | ···················· | ···················· | ···················· | ···················· | |
| M | 1771 hCoV-19/Wales/PHWC-47E7E6/2020\|EPI_ISL_595505\|2020 | ···················· | ···················· | ···················· | ···················· | ···················· | ···················· | |
| M | 1772 hCoV-19/Wales/PHWC-47E7F5/2020\|EPI_ISL_595506\|2020 | ···················· | ···················· | ···················· | ···················· | ···················· | ···················· | |
| M | 1774 hCoV-19/Wales/PHWC-47E82F/2020\|EPI_ISL_595509\|2020 | ···················· | ···················· | ···················· | ···················· | ···················· | ···················· | |
| M | 1775 hCoV-19/Wales/PHWC-47E86B/2020\|EPI_ISL_595512\|2020 | ···················· | ···················· | ···················· | ···················· | ···················· | ···················· | |
| M | 1776 hCoV-19/Wales/PHWC-47E87A/2020\|EPI_ISL_595513\|2020 | ···················· | ···················· | ···················· | ···················· | ···················· | ···················· | |
| M | 1777 hCoV-19/Wales/PHWC-47E889/2020\|EPI_ISL_595514\|2020 | ···················· | ···················· | ···················· | ···················· | ···················· | ···················· | |
| M | 1778 hCoV-19/Wales/PHWC-47E8A7/2020\|EPI_ISL_595515\|2020 | ···················· | ···················· | ···················· | ···················· | ···················· | ···················· | |
| M | 1779 hCoV-19/Wales/PHWC-47E8B6/2020\|EPI_ISL_595516\|2020 | ···················· | ···················· | ···················· | ···················· | NNN················· | ···················· | 1 |
| M | 1781 hCoV-19/Wales/PHWC-47E8D4/2020\|EPI_ISL_595518\|2020 | ···················· | ···················· | ···················· | ···················· | ···················· | ···················· | |
| M | 1782 hCoV-19/Wales/PHWC-47E8E3/2020\|EPI_ISL_595519\|2020 | ···················· | ···················· | ···················· | ···················· | ···················· | ···················· | |
| M | 1783 hCoV-19/Wales/PHWC-47E8F2/2020\|EPI_ISL_595520\|2020 | ···················· | ···················· | ···················· | ···················· | ···················· | ···················· | |
| M | 1784 hCoV-19/Wales/PHWC-47E94A/2020\|EPI_ISL_595525\|2020 | ···················· | ···················· | ···················· | ···················· | ···················· | ···················· | |
| M | 1785 hCoV-19/Wales/PHWC-47E959/2020\|EPI_ISL_595526\|2020 | ···················· | ···················· | ···················· | ···················· | ···················· | ···················· | |
| M | 1786 hCoV-19/Wales/PHWC-47E968/2020\|EPI_ISL_595527\|2020 | ···················· | ···················· | ···················· | ···················· | ···················· | ···················· | |
| M | 1787 hCoV-19/Wales/PHWC-47E986/2020\|EPI_ISL_595529\|2020 | ···················· | ···················· | ···················· | ···················· | ···················· | ···················· | |
| M | 1788 hCoV-19/Wales/PHWC-47E995/2020\|EPI_ISL_595530\|2020 | ···················· | ···················· | ···················· | ···················· | ···················· | ···················· | |
| M | 1789 hCoV-19/Scotland/QEUH-9F76F2/2020\|EPI_ISL_595696\|2 | ···················· | ···················· | ···················· | ···················· | ···················· | ···················· | |
| M | 1790 hCoV-19/Scotland/QEUH-9F771D/2020\|EPI_ISL_595697\|2 | ···················· | ···················· | ···················· | ···················· | ···················· | ···················· | |
| M | 1791 hCoV-19/England/QEUH-9F772C/2020\|EPI_ISL_595698\|20 | ···················· | ···················· | ···················· | ···················· | ···················· | ···················· | |
| M | 1792 hCoV-19/England/QEUH-9F773B/2020\|EPI_ISL_595699\|20 | ···················· | ···················· | ···················· | ···················· | ···················· | ···················· | |
| M | 1793 hCoV-19/England/ALDP-A2F4C9/2020\|EPI_ISL_587003\|20 | ···················· | ···················· | ···················· | ···················· | ···················· | ···················· | |
| M | 1794 hCoV-19/England/ALDP-A2EFE7/2020\|EPI_ISL_587006\|20 | ···················· | ···················· | ···················· | ···················· | ···················· | ···················· | |
| M | 1795 hCoV-19/England/ALDP-A2F915/2020\|EPI_ISL_587007\|20 | ···················· | ···················· | ···················· | ···················· | ···················· | ···················· | |
| M | 1796 hCoV-19/England/ALDP-A2FFC8/2020\|EPI_ISL_587008\|20 | ···················· | ···················· | ···················· | ···················· | ···················· | ···················· | |
| M | 1797 hCoV-19/England/ALDP-A2FD91/2020\|EPI_ISL_587009\|20 | ···················· | ···················· | ···················· | ···················· | ···················· | ···················· | |
| M | 1798 hCoV-19/England/ALDP-A2FDFB/2020\|EPI_ISL_587010\|20 | ···················· | ···················· | ···················· | ···················· | ···················· | ···················· | |
| M | 1799 hCoV-19/England/ALDP-A2EB98/2020\|EPI_ISL_587011\|20 | ···················· | ···················· | ···················· | ···················· | ···················· | ···················· | |
| M | 1800 hCoV-19/England/ALDP-A2EAB9/2020\|EPI_ISL_587012\|20 | ···················· | ···················· | ···················· | ···················· | ···················· | ···················· | |
| M | 1801 hCoV-19/England/ALDP-A3000D/2020\|EPI_ISL_587013\|20 | ···················· | ···················· | ···················· | ···················· | ···················· | ···················· | |
| M | 1802 hCoV-19/England/ALDP-A2F3BD/2020\|EPI_ISL_587014\|20 | ···················· | ···················· | ···················· | ···················· | ···················· | ···················· | |
| M | 1803 hCoV-19/England/ALDP-A2F04D/2020\|EPI_ISL_587015\|20 | ···················· | ···················· | ···················· | ···················· | ···················· | ···················· | |
| M | 1804 hCoV-19/England/ALDP-A2F4F6/2020\|EPI_ISL_587016\|20 | ···················· | ···················· | ···················· | ···················· | ···················· | ···················· | |
| M | 1805 hCoV-19/England/ALDP-A2ECFF/2020\|EPI_ISL_587039\|20 | ···················· | ···················· | ···················· | ···················· | ···················· | ···················· | |
| M | 1806 hCoV-19/England/ALDP-A2F353/2020\|EPI_ISL_587040\|20 | ···················· | ···················· | ···················· | ···················· | ···················· | ···················· | |
| M | 1807 hCoV-19/England/ALDP-A2FAF4/2020\|EPI_ISL_587041\|20 | ···················· | ···················· | ···················· | ···················· | ···················· | ···················· | |
| M | 1808 hCoV-19/England/ALDP-A2FDBF/2020\|EPI_ISL_587042\|20 | ···················· | ···················· | ···················· | ···················· | ···················· | ···················· | |
| M | 1809 hCoV-19/England/ALDP-A2EBA7/2020\|EPI_ISL_587043\|20 | ···················· | ···················· | ···················· | ···················· | ···················· | ···················· | |
| M | 1810 hCoV-19/England/ALDP-A2F5B7/2020\|EPI_ISL_587044\|20 | ···················· | ···················· | ···················· | ···················· | ···················· | ···················· | |
| M | 1811 hCoV-19/England/ALDP-A2EFF6/2020\|EPI_ISL_587045\|20 | ···················· | ···················· | ···················· | ···················· | ···················· | ···················· | |
| M | 1812 hCoV-19/England/ALDP-A2FC67/2020\|EPI_ISL_587047\|20 | ···················· | ···················· | ···················· | ···················· | ···················· | ···················· | |
| M | 1813 hCoV-19/England/ALDP-A2F344/2020\|EPI_ISL_587048\|20 | ···················· | ···················· | ···················· | ···················· | ···················· | ···················· | |
| M | 1814 hCoV-19/England/ALDP-A2F9AC/2020\|EPI_ISL_587049\|20 | ···················· | ···················· | ···················· | ···················· | ···················· | ···················· | |
| M | 1815 hCoV-19/England/ALDP-A2F98E/2020\|EPI_ISL_587050\|20 | ···················· | ···················· | ···················· | ···················· | ···················· | ···················· | |
| M | 1816 hCoV-19/England/ALDP-A2EDB0/2020\|EPI_ISL_587051\|20 | ···················· | ···················· | ···················· | ···················· | ···················· | ···················· | |
| M | 1817 hCoV-19/England/ALDP-A2FAA9/2020\|EPI_ISL_587052\|20 | ···················· | ···················· | ···················· | ···················· | ···················· | ···················· | |
| M | 1818 hCoV-19/England/ALDP-A2F4BA/2020\|EPI_ISL_587053\|20 | ···················· | ···················· | ···················· | ···················· | ···················· | ···················· | |
| M | 1819 hCoV-19/England/ALDP-A2F380/2020\|EPI_ISL_587054\|20 | ···················· | ···················· | ···················· | ···················· | ···················· | ···················· | |
| M | 1820 hCoV-19/England/ALDP-A2EFC9/2020\|EPI_ISL_587055\|20 | ···················· | ···················· | ···················· | ···················· | ···················· | ···················· | |
| M | 1821 hCoV-19/England/ALDP-A2F854/2020\|EPI_ISL_587056\|20 | ···················· | ···················· | ···················· | ···················· | ···················· | ···················· | |
| M | 1822 hCoV-19/England/ALDP-A2FCC1/2020\|EPI_ISL_587057\|20 | ···················· | ···················· | ···················· | ···················· | ···················· | ···················· | |
| M | 1823 hCoV-19/England/ALDP-A2F05C/2020\|EPI_ISL_587058\|20 | ···················· | ···················· | ···················· | ···················· | ···················· | ···················· | |
| M | 1824 hCoV-19/England/ALDP-A2FC3A/2020\|EPI_ISL_587059\|20 | ···················· | ···················· | ···················· | ···················· | ···················· | ···················· | |
| M | 1825 hCoV-19/England/ALDP-A2E952/2020\|EPI_ISL_587060\|20 | ···················· | ···················· | ···················· | ···················· | ···················· | ···················· | |
| M | 1826 hCoV-19/England/ALDP-A2FEAD/2020\|EPI_ISL_587061\|20 | ···················· | ···················· | ···················· | ···················· | ···················· | ···················· | |
| M | 1827 hCoV-19/England/ALDP-A2F265/2020\|EPI_ISL_587062\|20 | ···················· | ···················· | ···················· | ···················· | ···················· | ···················· | |
| M | 1828 hCoV-19/England/ALDP-A2EC59/2020\|EPI_ISL_587063\|20 | ···················· | ···················· | ···················· | ···················· | ···················· | ···················· | |
| M | 1829 hCoV-19/England/ALDP-A2F2A1/2020\|EPI_ISL_587064\|20 | ···················· | ···················· | ···················· | ···················· | ···················· | ···················· | |
| M | 1830 hCoV-19/England/ALDP-A2F7A2/2020\|EPI_ISL_587065\|20 | ···················· | ···················· | ···················· | ···················· | ···················· | ···················· | |
| M | 1831 hCoV-19/England/ALDP-A2EF7E/2020\|EPI_ISL_587066\|20 | ···················· | ···················· | ···················· | ···················· | ···················· | ···················· | |
| M | 1832 hCoV-19/England/ALDP-A2E9CB/2020\|EPI_ISL_587067\|20 | ···················· | ···················· | ···················· | ···················· | ···················· | ···················· | |
| M | 1833 hCoV-19/England/ALDP-A2F12C/2020\|EPI_ISL_587068\|20 | ···················· | ···················· | ···················· | ···················· | ···················· | ···················· | |
| M | 1834 hCoV-19/England/ALDP-A2F8AF/2020\|EPI_ISL_587069\|20 | ···················· | ···················· | ···················· | ···················· | ···················· | ···················· | |
| M | 1835 hCoV-19/England/ALDP-A2EA8C/2020\|EPI_ISL_587070\|20 | ···················· | ···················· | ···················· | ···················· | ···················· | ···················· | |
| M | 1836 hCoV-19/England/ALDP-A2EAC8/2020\|EPI_ISL_587071\|20 | ···················· | ···················· | ···················· | ···················· | ···················· | ···················· | |
| M | 1837 hCoV-19/Scotland/ALDP-A2F599/2020\|EPI_ISL_587072\|2 | ···················· | ···················· | ···················· | ···················· | ······T············· | ···················· | |
| M | 1838 hCoV-19/England/ALDP-A2EA6E/2020\|EPI_ISL_587073\|20 | ···················· | ···················· | ···················· | ···················· | ···················· | ···················· | |
| M | 1839 hCoV-19/England/ALDP-A2F9D9/2020\|EPI_ISL_587074\|20 | ···················· | ···················· | ···················· | ···················· | ···················· | ···················· | |

| | | | | | | | | |
|---|---|---|---|---|---|---|---|---|
| M | 1840 | hCoV-19/England/ALDP-A2FB2E/2020\|EPI_ISL_587075\|20 | ···················· | ···················· | ···················· | ···················· | ···················· | ···················· |
| M | 1841 | hCoV-19/England/ALDP-A2F10E/2020\|EPI_ISL_587076\|20 | ···················· | ···················· | ···················· | ···················· | ···················· | ···················· |
| M | 1842 | hCoV-19/England/ALDP-A2F7DF/2020\|EPI_ISL_587077\|20 | ···················· | ···················· | ···················· | ···················· | ···················· | ···················· |
| M | 1843 | hCoV-19/England/ALDP-A2FE61/2020\|EPI_ISL_587078\|20 | ···················· | ···················· | ···················· | ···················· | ···················· | ···················· |
| M | 1844 | hCoV-19/England/ALDP-A2F335/2020\|EPI_ISL_587079\|20 | ···················· | ···················· | ···················· | ···················· | ···················· | ···················· |
| M | 1845 | hCoV-19/England/ALDP-A2F02F/2020\|EPI_ISL_587100\|20 | ···················· | ···················· | ···················· | ···················· | ···················· | ···················· |
| M | 1846 | hCoV-19/England/ALDP-A2ED65/2020\|EPI_ISL_587101\|20 | ···················· | ···················· | ···················· | ···················· | ···················· | ···················· |
| M | 1847 | hCoV-19/England/ALDP-A2F54E/2020\|EPI_ISL_587102\|20 | ···················· | ···················· | ···················· | ···················· | ···················· | ···················· |
| M | 1848 | hCoV-19/England/ALDP-A2F97F/2020\|EPI_ISL_587103\|20 | ···················· | ···················· | ···················· | ···················· | ···················· | ···················· |
| M | 1849 | hCoV-19/England/ALDP-A2F933/2020\|EPI_ISL_587104\|20 | ···················· | ···················· | ···················· | ···················· | ···················· | ···················· |
| M | 1850 | hCoV-19/England/ALDP-A2FA12/2020\|EPI_ISL_587105\|20 | ···················· | ···················· | ···················· | ···················· | ···················· | ···················· |
| M | 1851 | hCoV-19/England/ALDP-A2EEEA/2020\|EPI_ISL_587106\|20 | ···················· | ···················· | ···················· | ···················· | ···················· | ···················· |
| M | 1852 | hCoV-19/England/ALDP-A2F9E8/2020\|EPI_ISL_587107\|20 | ···················· | ···················· | ···················· | ···················· | ···················· | ···················· |
| M | 1853 | hCoV-19/England/ALDP-A2F890/2020\|EPI_ISL_587108\|20 | ···················· | ···················· | ···················· | ···················· | ···················· | ···················· |
| M | 1854 | hCoV-19/England/ALDP-A2F1A4/2020\|EPI_ISL_587109\|20 | ···················· | ···················· | ···················· | ···················· | ···················· | ···················· |
| M | 1855 | hCoV-19/England/ALDP-A2F8EB/2020\|EPI_ISL_587110\|20 | ···················· | ·A·················· | ···················· | ···················· | ···················· | ···A················ |
| M | 1856 | hCoV-19/England/ALDP-A2FA8B/2020\|EPI_ISL_587111\|20 | ···················· | ···················· | ···················· | ···················· | ···················· | ···················· |
| M | 1857 | hCoV-19/England/QEUH-9F47A7/2020\|EPI_ISL_588333\|20 | ···················· | ···················· | ···················· | ···················· | ···················· | ···················· |
| M | 1858 | hCoV-19/England/QEUH-9F5B7D/2020\|EPI_ISL_588334\|20 | ···················· | ···················· | ···················· | ···················· | ······T············· | ···················· |
| M | 1859 | hCoV-19/England/QEUH-9F48C2/2020\|EPI_ISL_588337\|20 | ···················· | ···················· | ···················· | ···················· | ···················· | ···················· |
| M | 1860 | hCoV-19/England/QEUH-9F5C5C/2020\|EPI_ISL_588338\|20 | ···················· | ···················· | ···················· | ···················· | ···················· | ···················· |
| M | 1861 | hCoV-19/England/QEUH-9F68A2/2020\|EPI_ISL_588339\|20 | ···················· | ·A·················· | ···················· | ···················· | ···················· | ···A················ |
| M | 1862 | hCoV-19/England/QEUH-9F6918/2020\|EPI_ISL_588340\|20 | ···················· | ···················· | ···················· | ···················· | ···················· | ···················· |
| M | 1863 | hCoV-19/England/QEUH-9F5ACB/2020\|EPI_ISL_588341\|20 | ···················· | ···················· | ···················· | ···················· | ···················· | ···················· |
| M | 1864 | hCoV-19/England/QEUH-9F490B/2020\|EPI_ISL_588342\|20 | ···················· | ·A·················· | ···················· | ···················· | ···················· | ···A················ |
| M | 1865 | hCoV-19/England/QEUH-9F5014/2020\|EPI_ISL_588343\|20 | ···················· | ···················· | ···················· | ···················· | ···················· | ···················· |
| M | 1866 | hCoV-19/England/QEUH-9F6A9D/2020\|EPI_ISL_588344\|20 | ···················· | ···················· | ···················· | ···················· | ···················· | ···················· |
| M | 1867 | hCoV-19/England/QEUH-9F5849/2020\|EPI_ISL_588345\|20 | ···················· | ···················· | ···················· | ···················· | ····T··············· | ···················· |
| M | 1868 | hCoV-19/England/QEUH-9F4BD8/2020\|EPI_ISL_588346\|20 | ···················· | ···················· | ···················· | ···················· | ···················· | ···················· |
| M | 1869 | hCoV-19/England/QEUH-9F482C/2020\|EPI_ISL_588370\|20 | ···················· | ···················· | ···················· | ···················· | ···················· | ···················· |
| M | 1870 | hCoV-19/England/QEUH-9F58FE/2020\|EPI_ISL_588371\|20 | ···················· | ···················· | ···················· | ···················· | ···················· | ···················· |
| M | 1871 | hCoV-19/England/QEUH-9F5C01/2020\|EPI_ISL_588372\|20 | ···················· | ···················· | ···················· | ···················· | ···················· | ···················· |
| M | 1872 | hCoV-19/England/QEUH-9F4482/2020\|EPI_ISL_588374\|20 | ···················· | ···················· | ···················· | ···················· | ······T············· | ···················· |
| M | 1873 | hCoV-19/England/QEUH-9F4FFA/2020\|EPI_ISL_588375\|20 | ···················· | ···················· | ···················· | ···················· | ···················· | ···················· |
| M | 1874 | hCoV-19/England/QEUH-9F4ACC/2020\|EPI_ISL_588376\|20 | ···················· | ···················· | ···················· | ···················· | ···················· | ···················· |
| M | 1875 | hCoV-19/England/QEUH-9F5A61/2020\|EPI_ISL_588377\|20 | ···················· | ···················· | ···················· | ···················· | ···················· | ···················· |
| M | 1876 | hCoV-19/England/QEUH-9F51D5/2020\|EPI_ISL_588378\|20 | ···················· | ···················· | ···················· | ···················· | ···················· | ···················· |
| M | 1877 | hCoV-19/England/QEUH-9F5199/2020\|EPI_ISL_588379\|20 | ···················· | ···················· | ···················· | ···················· | ···················· | ···················· |
| M | 1878 | hCoV-19/England/QEUH-9F5C98/2020\|EPI_ISL_588381\|20 | ···················· | ···················· | ···················· | ···················· | ···················· | ···················· |
| M | 1879 | hCoV-19/England/QEUH-9F5ADA/2020\|EPI_ISL_588382\|20 | ···················· | ···················· | ···················· | ···················· | ···················· | ···················· |
| M | 1880 | hCoV-19/England/QEUH-9F69DC/2020\|EPI_ISL_588383\|20 | ···················· | ···················· | ···················· | ···················· | ···················· | ···················· |
| M | 1881 | hCoV-19/England/QEUH-9F49DE/2020\|EPI_ISL_588384\|20 | ···················· | ···················· | ···················· | ···················· | ···················· | ···················· |
| M | 1882 | hCoV-19/England/QEUH-9F4D5A/2020\|EPI_ISL_588385\|20 | ···················· | ···················· | ···················· | ···················· | ···················· | ···················· |
| M | 1883 | hCoV-19/England/QEUH-9F4895/2020\|EPI_ISL_588386\|20 | ···················· | ···················· | ···················· | ···················· | ···················· | ···················· |
| M | 1884 | hCoV-19/England/QEUH-9F4868/2020\|EPI_ISL_588387\|20 | ···················· | ···················· | ···················· | ···················· | ···················· | ···················· |
| M | 1885 | hCoV-19/England/QEUH-9F5B13/2020\|EPI_ISL_588388\|20 | ···················· | ···················· | ···················· | ···················· | ···················· | ···················· |
| M | 1886 | hCoV-19/England/QEUH-9F64F9/2020\|EPI_ISL_588389\|20 | ···················· | ···················· | ···················· | ···················· | ···················· | ···················· |
| M | 1887 | hCoV-19/England/QEUH-9F5B5F/2020\|EPI_ISL_588391\|20 | ···················· | ···················· | ···················· | ···················· | ···················· | ···················· |
| M | 1888 | hCoV-19/England/QEUH-9F663F/2020\|EPI_ISL_588392\|20 | ···················· | ·A·················· | ···················· | ···················· | ···················· | ···A················ |
| M | 1889 | hCoV-19/England/QEUH-9F6620/2020\|EPI_ISL_588393\|20 | ···················· | ·A·················· | ···················· | ···················· | ···················· | ···A················ |
| M | 1890 | hCoV-19/England/QEUH-9F4D96/2020\|EPI_ISL_588394\|20 | ···················· | ···················· | ···················· | ···················· | ···················· | ···················· |
| M | 1891 | hCoV-19/England/QEUH-9F5269/2020\|EPI_ISL_588395\|20 | ···················· | ···················· | ···················· | ···················· | ···················· | ···················· |
| M | 1892 | hCoV-19/England/QEUH-9F49A1/2020\|EPI_ISL_588397\|20 | ···················· | ···················· | ···················· | ···················· | ···················· | ···················· |
| M | 1893 | hCoV-19/England/QEUH-9F5D86/2020\|EPI_ISL_588398\|20 | ···················· | ···················· | ···················· | ···················· | ···················· | ···················· |
| M | 1894 | hCoV-19/England/QEUH-9F475C/2020\|EPI_ISL_588399\|20 | ···················· | ·A·················· | ···················· | ···················· | ···················· | ···A················ |
| M | 1895 | hCoV-19/England/QEUH-9F5FEA/2020\|EPI_ISL_588400\|20 | ···················· | ···················· | ···················· | ···················· | ···················· | ···················· |
| M | 1896 | hCoV-19/England/QEUH-9F4983/2020\|EPI_ISL_588402\|20 | ···················· | ···················· | ···················· | ···················· | ···················· | ···················· |
| M | 1897 | hCoV-19/England/QEUH-9F5CB6/2020\|EPI_ISL_588425\|20 | ···················· | ···················· | ···················· | ···················· | ···················· | ···················· |
| M | 1898 | hCoV-19/England/QEUH-9F4E84/2020\|EPI_ISL_588426\|20 | ···················· | ···················· | ···················· | ···················· | ···················· | ···················· |
| M | 1899 | hCoV-19/England/QEUH-9F6A24/2020\|EPI_ISL_588427\|20 | ···················· | ···················· | ···················· | ···················· | ···················· | ···················· |
| M | 1900 | hCoV-19/England/QEUH-9F4B6F/2020\|EPI_ISL_588428\|20 | ···················· | ···················· | ···················· | ···················· | ···················· | ···················· |
| M | 1901 | hCoV-19/England/QEUH-9F581C/2020\|EPI_ISL_588429\|20 | ···················· | ···················· | ···················· | ···················· | ···················· | ···················· |
| M | 1902 | hCoV-19/England/QEUH-9F6514/2020\|EPI_ISL_588430\|20 | ···················· | ···················· | ···················· | ···················· | ···················· | ···················· |
| M | 1903 | hCoV-19/England/QEUH-9F4938/2020\|EPI_ISL_588431\|20 | ···················· | ···················· | ···················· | ···················· | ···················· | ···················· |
| M | 1904 | hCoV-19/England/QEUH-9F5A52/2020\|EPI_ISL_588432\|20 | ···················· | ···················· | ···················· | ···················· | ···················· | ···················· |
| M | 1905 | hCoV-19/England/QEUH-9F58C1/2020\|EPI_ISL_588433\|20 | ···················· | ·A·················· | ···················· | ···················· | ···················· | ···A················ |
| M | 1906 | hCoV-19/England/QEUH-9F68DF/2020\|EPI_ISL_588434\|20 | ···················· | ···················· | ···················· | ···················· | ···················· | ···················· |
| M | 1907 | hCoV-19/England/QEUH-9F5B40/2020\|EPI_ISL_588435\|20 | ···················· | ·A·················· | ···················· | ···················· | ···················· | ···A················ |
| M | 1908 | hCoV-19/England/QEUH-9F5BC8/2020\|EPI_ISL_588436\|20 | ···················· | ···················· | ···················· | ···················· | ···················· | ···················· |
| M | 1909 | hCoV-19/England/QEUH-9F5C6B/2020\|EPI_ISL_588437\|20 | ···················· | ·A·················· | ···················· | ···················· | ···················· | ···················· |
| M | 1910 | hCoV-19/England/QEUH-9F4B50/2020\|EPI_ISL_588439\|20 | ···················· | ···················· | ···················· | ···················· | ···················· | ···················· |
| M | 1911 | hCoV-19/England/QEUH-9F4EC0/2020\|EPI_ISL_588441\|20 | ···················· | ···················· | ···················· | ···················· | ···················· | ···················· |
| M | 1912 | hCoV-19/England/QEUH-9F665D/2020\|EPI_ISL_588442\|20 | ···················· | ···················· | ···················· | ···················· | ···················· | ···················· |
| M | 1913 | hCoV-19/England/MILK-9F187A/2020\|EPI_ISL_589664\|20 | ···················· | ···················· | ···················· | ···················· | ···················· | ···················· |
| M | 1914 | hCoV-19/Wales/MILK-9F1670/2020\|EPI_ISL_589668\|2020 | ···················· | ···················· | ···················· | ···················· | ···················· | ···················· |
| M | 1915 | hCoV-19/England/MILK-9F1607/2020\|EPI_ISL_589669\|20 | ···················· | ···················· | ···················· | ···················· | ···················· | ···················· |
| M | 1916 | hCoV-19/England/MILK-9F1388/2020\|EPI_ISL_589670\|20 | ···················· | ···················· | ···················· | ···················· | ···················· | ···················· |
| M | 1917 | hCoV-19/England/MILK-9F19C2/2020\|EPI_ISL_589671\|20 | ···················· | ·A·················· | ···················· | ···················· | ···················· | ···A················ |
| M | 1918 | hCoV-19/England/MILK-9F08D5/2020\|EPI_ISL_589672\|20 | ···················· | ·A·················· | ···················· | ···················· | ···················· | ···A················ |
| M | 1919 | hCoV-19/England/MILK-9F133D/2020\|EPI_ISL_589673\|20 | ···················· | ···················· | ···················· | ···················· | ···················· | ···················· |

| | | | | | | | | |
|---|---|---|---|---|---|---|---|---|
| M | 1920 | hCoV-19/England/MILK-9F1EC3/2020\|EPI_ISL_589674\|20 | ···················· | ···················· | ···················· | ···················· | ···················· | ···················· |
| M | 1921 | hCoV-19/England/MILK-9F1977/2020\|EPI_ISL_589675\|20 | ···················· | ···················· | ···················· | ···················· | ···················· | ···················· |
| M | 1922 | hCoV-19/England/MILK-9F1106/2020\|EPI_ISL_589700\|20 | ···················· | ···················· | ···················· | ···················· | ···················· | ···················· |
| M | 1923 | hCoV-19/England/MILK-9F1CBA/2020\|EPI_ISL_589702\|20 | ···················· | ···················· | ···················· | ···················· | ···················· | ···················· |
| M | 1924 | hCoV-19/England/MILK-9F15EC/2020\|EPI_ISL_589703\|20 | ···················· | ···················· | ···················· | ···················· | ···················· | ···················· |
| M | 1925 | hCoV-19/England/MILK-9F0996/2020\|EPI_ISL_589705\|20 | ···················· | ···················· | ···················· | ···················· | ···················· | ···················· |
| M | 1926 | hCoV-19/England/MILK-9F0AFD/2020\|EPI_ISL_589706\|20 | ···················· | ···················· | ···················· | ···················· | ···················· | ···················· |
| M | 1927 | hCoV-19/England/MILK-9F0AB1/2020\|EPI_ISL_589708\|20 | ···················· | ···················· | ···················· | ···················· | ···················· | ···················· |
| M | 1928 | hCoV-19/England/MILK-9F119D/2020\|EPI_ISL_589709\|20 | ···················· | ···················· | ···················· | ···················· | ···················· | ···················· |
| M | 1929 | hCoV-19/England/MILK-9F1C23/2020\|EPI_ISL_589711\|20 | ···················· | ···················· | ···················· | ···················· | ···················· | ···················· |
| M | 1930 | hCoV-19/England/MILK-9F0A93/2020\|EPI_ISL_589712\|20 | ···················· | ···················· | ···················· | ···················· | ···················· | ···················· |
| M | 1931 | hCoV-19/England/MILK-9F1CF6/2020\|EPI_ISL_589714\|20 | ···················· | ···················· | ···················· | ···················· | ···················· | ···················· |
| M | 1932 | hCoV-19/Wales/MILK-9F1E69/2020\|EPI_ISL_589715\|2020 | ···················· | ···················· | ···················· | ···················· | ···················· | ···················· |
| M | 1933 | hCoV-19/England/MILK-9F18F2/2020\|EPI_ISL_589716\|20 | ···················· | ···················· | ···················· | ···················· | ···················· | ···················· |
| M | 1934 | hCoV-19/England/MILK-9F1968/2020\|EPI_ISL_589717\|20 | ···················· | ·A·················· | ···················· | ···················· | ···················· | ···A················ |
| M | 1935 | hCoV-19/England/MILK-9F14FE/2020\|EPI_ISL_589718\|20 | ···················· | ···················· | ···················· | ···················· | ···················· | ···················· |
| M | 1936 | hCoV-19/England/MILK-9F0CBB/2020\|EPI_ISL_589719\|20 | ···················· | ···················· | ···················· | ···················· | ···················· | ···················· |
| M | 1937 | hCoV-19/England/MILK-9F2017/2020\|EPI_ISL_589720\|20 | ···················· | ···················· | ···················· | ···················· | ······T············· | ···················· |
| M | 1938 | hCoV-19/England/MILK-9F090F/2020\|EPI_ISL_589721\|20 | ···················· | ···················· | ···················· | ···················· | ···················· | ···················· |
| M | 1939 | hCoV-19/England/MILK-9F0BCD/2020\|EPI_ISL_589723\|20 | ···················· | ···················· | ···················· | ···················· | ···················· | ···················· |
| M | 1940 | hCoV-19/Wales/MILK-9F0EE2/2020\|EPI_ISL_589724\|2020 | ···················· | ···················· | ···················· | ···················· | ···················· | ···················· |
| M | 1941 | hCoV-19/England/MILK-9F09E1/2020\|EPI_ISL_589725\|20 | ···················· | ···················· | ···················· | ···················· | ···················· | ···················· |
| M | 1942 | hCoV-19/England/MILK-9EF578/2020\|EPI_ISL_589726\|20 | ···················· | ···················· | ···················· | ···················· | ···················· | ···················· |
| M | 1943 | hCoV-19/England/MILK-9F1DD5/2020\|EPI_ISL_589727\|20 | ···················· | ···················· | ···················· | ···················· | ···················· | ···················· |
| M | 1944 | hCoV-19/England/MILK-9F1F39/2020\|EPI_ISL_589728\|20 | ···················· | ···················· | ···················· | ···················· | ···················· | ···················· |
| M | 1945 | hCoV-19/England/MILK-9F0B45/2020\|EPI_ISL_589729\|20 | ···················· | ···················· | ···················· | ···················· | ···················· | ···················· |
| M | 1946 | hCoV-19/England/MILK-9F1889/2020\|EPI_ISL_589730\|20 | ···················· | ···················· | ···················· | ···················· | ···················· | ···················· |
| M | 1947 | hCoV-19/England/MILK-9F1BAE/2020\|EPI_ISL_589731\|20 | ···················· | ···················· | ···················· | ···················· | ···················· | ···················· |
| M | 1948 | hCoV-19/England/MILK-9F2035/2020\|EPI_ISL_589732\|20 | ···················· | ·A·················· | ···················· | ···················· | ···················· | ···A················ |
| M | 1949 | hCoV-19/England/MILK-9F19D1/2020\|EPI_ISL_589757\|20 | ···················· | ···················· | ···················· | ···················· | ···················· | ···················· |
| M | 1950 | hCoV-19/England/MILK-9EF5E1/2020\|EPI_ISL_589759\|20 | ···················· | ···················· | ···················· | ···················· | ···················· | ···················· |
| M | 1951 | hCoV-19/England/MILK-9F0D6D/2020\|EPI_ISL_589760\|20 | ···················· | ···················· | ···················· | ···················· | ···················· | ···················· |
| M | 1952 | hCoV-19/England/MILK-9F085D/2020\|EPI_ISL_589761\|20 | ···················· | ···················· | ···················· | ···················· | ···················· | ···················· |
| M | 1953 | hCoV-19/England/ALDP-9E609E/2020\|EPI_ISL_589762\|20 | ···················· | ···················· | ···················· | ···················· | ···················· | ···················· |
| M | 1954 | hCoV-19/Wales/MILK-9F0240/2020\|EPI_ISL_589763\|2020 | ···················· | ···················· | ···················· | ···················· | ···················· | ···················· |
| M | 1955 | hCoV-19/Wales/MILK-9F0231/2020\|EPI_ISL_589764\|2020 | ···················· | ···················· | ···················· | ···················· | ···················· | ···················· |
| M | 1956 | hCoV-19/England/ALDP-9EDD63/2020\|EPI_ISL_589765\|20 | ···················· | ···················· | ···················· | ···················· | ···················· | ···················· |
| M | 1957 | hCoV-19/Wales/MILK-9F032F/2020\|EPI_ISL_589766\|2020 | ···················· | ···················· | ···················· | ···················· | ···················· | ···················· |
| M | 1958 | hCoV-19/Wales/MILK-9EEB77/2020\|EPI_ISL_589770\|2020 | ···················· | ···················· | ···················· | ···················· | ···················· | ···················· |
| M | 1959 | hCoV-19/Wales/MILK-9F025F/2020\|EPI_ISL_589771\|2020 | ···················· | ···················· | ···················· | ···················· | ···················· | ···················· |
| M | 1960 | hCoV-19/Wales/MILK-9EE61C/2020\|EPI_ISL_589772\|2020 | ···················· | ···················· | ···················· | ···················· | ···················· | ···················· |
| M | 1961 | hCoV-19/Wales/MILK-9F0222/2020\|EPI_ISL_589773\|2020 | ···················· | ···················· | ···················· | ···················· | ···················· | ···················· |
| M | 1962 | hCoV-19/England/QEUH-9B7667/2020\|EPI_ISL_590645\|20 | ···················· | ···················· | ···················· | ···················· | ···················· | ···················· |
| M | 1963 | hCoV-19/England/QEUH-9B71A2/2020\|EPI_ISL_590646\|20 | ···················· | ···················· | ···················· | ···················· | ···················· | ···················· |
| M | 1964 | hCoV-19/England/QEUH-9B8129/2020\|EPI_ISL_590647\|20 | ···················· | ···················· | ···················· | ···················· | ···················· | ···················· |
| M | 1965 | hCoV-19/England/QEUH-9B795F/2020\|EPI_ISL_590648\|20 | ···················· | ···················· | ···················· | ···················· | ···················· | ···················· |
| M | 1966 | hCoV-19/England/QEUH-9B7281/2020\|EPI_ISL_590649\|20 | ···················· | ···················· | ···················· | ···················· | ···················· | ···················· |
| M | 1967 | hCoV-19/England/QEUH-9B804A/2020\|EPI_ISL_590650\|20 | ···················· | ···················· | ···················· | ···················· | ···················· | ···················· |
| M | 1968 | hCoV-19/England/QEUH-9B788F/2020\|EPI_ISL_590651\|20 | ···················· | ···················· | ···················· | ···················· | ···················· | ···················· |
| M | 1969 | hCoV-19/England/QEUH-9B78BC/2020\|EPI_ISL_590652\|20 | ···················· | ···················· | ···················· | ···················· | ···················· | ···················· |
| M | 1970 | hCoV-19/England/QEUH-9B7694/2020\|EPI_ISL_590654\|20 | ···················· | ···················· | ···················· | ···················· | ···················· | ···················· |
| M | 1971 | hCoV-19/England/QEUH-9B6F8B/2020\|EPI_ISL_590655\|20 | ···················· | ···················· | ···················· | ···················· | ···················· | ···················· |
| M | 1972 | hCoV-19/England/QEUH-9B7DF9/2020\|EPI_ISL_590656\|20 | ···················· | ···················· | ···················· | ···················· | ···················· | ···················· |
| M | 1973 | hCoV-19/England/QEUH-9B7A98/2020\|EPI_ISL_590657\|20 | ···················· | ···················· | ···················· | ···················· | ···················· | ···················· |
| M | 1974 | hCoV-19/England/QEUH-9B7EC9/2020\|EPI_ISL_590658\|20 | ···················· | ···················· | ···················· | ···················· | ···················· | ···················· |
| M | 1975 | hCoV-19/England/QEUH-9B7A3E/2020\|EPI_ISL_590681\|20 | ···················· | ···················· | ···················· | ···················· | ···················· | ···················· |
| M | 1976 | hCoV-19/England/QEUH-9B7C0B/2020\|EPI_ISL_590682\|20 | ···················· | ···················· | ···················· | ···················· | ···················· | ···················· |
| M | 1977 | hCoV-19/England/QEUH-9B7852/2020\|EPI_ISL_590683\|20 | ···················· | ···················· | ···················· | ···················· | ····T··············· | ···················· |
| M | 1978 | hCoV-19/England/QEUH-9B6FB8/2020\|EPI_ISL_590684\|20 | ···················· | ···················· | ···················· | ···················· | ···················· | ···················· |
| M | 1979 | hCoV-19/England/QEUH-9B7E41/2020\|EPI_ISL_590685\|20 | ···················· | ···················· | ···················· | ···················· | ···················· | ···················· |
| M | 1980 | hCoV-19/England/QEUH-9B7834/2020\|EPI_ISL_590686\|20 | ···················· | ···················· | ···················· | ···················· | ····T··············· | ···················· |
| M | 1981 | hCoV-19/England/QEUH-9B7290/2020\|EPI_ISL_590687\|20 | ···················· | ···················· | ···················· | ···················· | ···················· | ···················· |
| M | 1982 | hCoV-19/England/QEUH-9B7A7A/2020\|EPI_ISL_590688\|20 | ···················· | ···················· | ···················· | ···················· | ···················· | ···················· |
| M | 1983 | hCoV-19/Italy/LAZ-INMI-25/2020\|EPI_ISL_590693\|2020 | ···················· | ···················· | ···················· | ···················· | ···················· | ···················· |
| M | 1984 | hCoV-19/Italy/LAZ-INMI-26/2020\|EPI_ISL_590694\|2020 | ···················· | ···················· | ···················· | ···················· | ···················· | ···················· |
| M | 1985 | hCoV-19/Italy/LAZ-INMI-27/2020\|EPI_ISL_590695\|2020 | ···················· | ···················· | ···················· | ···················· | ···················· | ···················· |
| M | 1986 | hCoV-19/Italy/LAZ-INMI-15/2020\|EPI_ISL_590696\|2020 | ···················· | ···················· | ···················· | ···················· | ···················· | ···················· |
| M | 1987 | hCoV-19/Italy/LAZ-INMI-28/2020\|EPI_ISL_590697\|2020 | ···················· | ···················· | ···················· | ···················· | ···················· | ···················· |
| M | 1988 | hCoV-19/Italy/LAZ-INMI-29/2020\|EPI_ISL_590698\|2020 | ···················· | ···················· | ···················· | ···················· | ···················· | ···················· |
| M | 1989 | hCoV-19/USA/MI-UM-10036139552/2020\|EPI_ISL_590699\| | ···················· | ···················· | ···················· | ···················· | ···················· | ···················· |
| M | 1990 | hCoV-19/USA/MI-UM-10036112787/2020\|EPI_ISL_590700\| | ···················· | ···················· | ···················· | ···················· | ···················· | ···················· |
| M | 1991 | hCoV-19/USA/MI-UM-10036119854/2020\|EPI_ISL_590701\| | ···················· | ···················· | ···················· | ···················· | ···················· | ···················· |
| M | 1992 | hCoV-19/USA/MI-UM-10036127680/2020\|EPI_ISL_590702\| | ···················· | ···················· | ···················· | ···················· | ···················· | ···················· |
| M | 1993 | hCoV-19/USA/MI-UM-10036116407/2020\|EPI_ISL_590703\| | ···················· | ···················· | ···················· | ···················· | ···················· | ···················· |
| M | 1994 | hCoV-19/USA/MI-UM-10036130933/2020\|EPI_ISL_590704\| | ···················· | ···················· | ···················· | ···················· | ···················· | ···················· |
| M | 1995 | hCoV-19/USA/MI-UM-10036139449/2020\|EPI_ISL_590705\| | ···················· | ···················· | ···················· | ···················· | ···················· | ···················· |
| M | 1996 | hCoV-19/USA/MI-UM-10036117092/2020\|EPI_ISL_590706\| | ···················· | ···················· | ···················· | ···················· | ···················· | ···················· |
| M | 1997 | hCoV-19/USA/MI-UM-10036147927/2020\|EPI_ISL_590707\| | ···················· | ·A·················· | ···················· | ···················· | ···················· | ···················· |
| M | 1998 | hCoV-19/USA/MI-UM-10036114941/2020\|EPI_ISL_590708\| | ···················· | ···················· | ···················· | ···················· | ···················· | ···················· |
| M | 1999 | hCoV-19/USA/MI-UM-10036133006/2020\|EPI_ISL_590709\| | ···················· | ·············A······ | ···················· | ···················· | ···················· | ···················· |

| | | | | | | | |
|---|---|---|---|---|---|---|---|
| M | 2000 hCoV-19/USA/MI-UM-10036139493/2020\|EPI_ISL_590710\| | .................... | .................... | .................... | .................... | .................... | .................... |
| M | 2001 hCoV-19/USA/MI-UM-10036138820/2020\|EPI_ISL_590711\| | .................... | .................... | .................... | .................... | .................... | .................... |
| M | 2002 hCoV-19/USA/MI-UM-10036115045/2020\|EPI_ISL_590712\| | .................... | .................... | .................... | .................... | .................... | .................... |
| M | 2003 hCoV-19/USA/MI-UM-10036099219/2020\|EPI_ISL_590713\| | .................... | .............A...... | .................... | .................... | .................... | .................... |
| M | 2005 hCoV-19/USA/MI-UM-10036132232/2020\|EPI_ISL_590715\| | .................... | .............A...... | .................... | .................... | .................... | .................... |
| M | 2006 hCoV-19/USA/MI-UM-10036132857/2020\|EPI_ISL_590716\| | .................... | .............A...... | .................... | .................... | .................... | .................... |
| M | 2007 hCoV-19/USA/MI-UM-10036117605/2020\|EPI_ISL_590717\| | .................... | .................... | .................... | .................... | .................... | .................... |
| M | 2008 hCoV-19/USA/MI-UM-10036119783/2020\|EPI_ISL_590718\| | .................... | .................... | .................... | .................... | .................... | .................... |
| M | 2009 hCoV-19/USA/MI-UM-10036128032/2020\|EPI_ISL_590719\| | .................... | .................... | .................... | .................... | .................... | .................... |
| M | 2011 hCoV-19/USA/MI-UM-10036255029/2020\|EPI_ISL_590741\| | .................... | .................... | .................... | .................... | .................... | .................... |
| M | 2012 hCoV-19/USA/MI-UM-10036212950/2020\|EPI_ISL_590742\| | .................... | .................... | .................... | .................... | .................... | .................... |
| M | 2013 hCoV-19/USA/MI-UM-10036207074/2020\|EPI_ISL_590743\| | .................... | .................... | .................... | .................... | .................... | .................... |
| M | 2014 hCoV-19/USA/MI-UM-10036210275/2020\|EPI_ISL_590744\| | .................... | .................... | .................... | .................... | .................... | .................... |
| M | 2015 hCoV-19/USA/MI-UM-10036187521/2020\|EPI_ISL_590745\| | ...........T........ | .............A...... | .................... | .................... | .................... | .................... |
| M | 2016 hCoV-19/USA/MI-UM-10036182091/2020\|EPI_ISL_590746\| | .................... | .................... | .................... | .................... | .................... | .................... |
| M | 2017 hCoV-19/USA/MI-UM-MH27428/2020\|EPI_ISL_590747\|2020 | .................... | .................... | .................... | .................... | .................... | .................... |
| M | 2018 hCoV-19/USA/MI-UM-MH27455/2020\|EPI_ISL_590748\|2020 | .................... | .............A...... | .................... | .................... | .................... | .................... |
| M | 2019 hCoV-19/USA/MI-UM-MH27501/2020\|EPI_ISL_590749\|2020 | .................... | .................... | .................... | .................... | .................... | .................... |
| M | 2020 hCoV-19/USA/MI-UM-MH27525/2020\|EPI_ISL_590750\|2020 | .................... | .................... | .................... | .................... | .................... | .................... |
| M | 2022 hCoV-19/USA/MN-MDH-1748/2020\|EPI_ISL_590753\|2020-0 | .................... | .................... | .................... | .................... | .................... | .................... |
| M | 2023 hCoV-19/Australia/VIC12374/2020\|EPI_ISL_591970\|202 | .................... | .................... | .................... | .................... | .................... | .................... |
| M | 2024 hCoV-19/Australia/VIC12388/2020\|EPI_ISL_591975\|202 | .................... | .................... | .................... | .................... | .................... | .................... |
| M | 2025 hCoV-19/Australia/VIC12395/2020\|EPI_ISL_591976\|202 | .................... | .................... | .................... | .................... | .................... | .................... |
| M | 2026 hCoV-19/Australia/VIC12399/2020\|EPI_ISL_591977\|202 | .................... | .................... | .................... | .................... | NNN................. | .................... 1 |
| M | 2027 hCoV-19/Australia/VIC12404/2020\|EPI_ISL_591978\|202 | .................... | .................... | .................... | .................... | .................... | .................... |
| M | 2028 hCoV-19/Australia/VIC12408/2020\|EPI_ISL_591979\|202 | .................... | .................... | .................... | .................... | .................... | .................... |
| M | 2029 hCoV-19/Australia/VIC12409/2020\|EPI_ISL_591980\|202 | .................... | .................... | .................... | .................... | .................... | .................... |
| M | 2030 hCoV-19/Australia/VIC12413/2020\|EPI_ISL_591981\|202 | .................... | .................... | .................... | .................... | .................... | .................... |
| M | 2031 hCoV-19/Australia/VIC12417/2020\|EPI_ISL_591982\|202 | .................... | .................... | .................... | .................... | .................... | .................... |
| M | 2032 hCoV-19/Australia/VIC12419/2020\|EPI_ISL_591983\|202 | .................... | .................... | .................... | .................... | .................... | .................... |
| M | 2034 hCoV-19/Australia/VIC12424/2020\|EPI_ISL_591985\|202 | .................... | .................... | .................... | .................... | .................... | .................... |
| M | 2035 hCoV-19/Australia/VIC12427/2020\|EPI_ISL_591986\|202 | .................... | .................... | .................... | .................... | .................... | .................... |
| M | 2037 hCoV-19/Australia/VIC12430/2020\|EPI_ISL_591988\|202 | .................... | .................... | .................... | .................... | .................... | .................... |
| M | 2038 hCoV-19/Australia/VIC12431/2020\|EPI_ISL_591989\|202 | .................... | .................... | .................... | .................... | .................... | .................... |
| M | 2039 hCoV-19/USA/FL-BPHL-1598/2020\|EPI_ISL_594288\|2020- | .................... | .................... | .................... | .................... | .................... | .................... |
| M | 2040 hCoV-19/USA/FL-BPHL-1599/2020\|EPI_ISL_594289\|2020- | .................... | .................... | .................... | .................... | .................... | .................... |
| M | 2041 hCoV-19/USA/FL-BPHL-1600/2020\|EPI_ISL_594290\|2020- | .................... | .................... | .................... | .................... | .................... | .................... |
| M | 2043 hCoV-19/USA/FL-BPHL-1603/2020\|EPI_ISL_594293\|2020- | .................... | .................... | .................... | .................... | .................... | .................... |
| M | 2044 hCoV-19/USA/FL-BPHL-1604/2020\|EPI_ISL_594294\|2020- | .................... | .................... | .................... | .................... | .................... | .................... |
| M | 2046 hCoV-19/USA/FL-BPHL-1606/2020\|EPI_ISL_594296\|2020- | .................... | .................... | .................... | .................... | .................... | .................... |
| M | 2047 hCoV-19/USA/FL-BPHL-1607/2020\|EPI_ISL_594297\|2020- | .................... | .................... | .................... | .................... | .................... | .................... |
| M | 2048 hCoV-19/USA/FL-BPHL-1609/2020\|EPI_ISL_594299\|2020- | .................... | .................... | .................... | .................... | .................... | .................... |
| M | 2049 hCoV-19/Austria/CeMM0057/2020\|EPI_ISL_583557\|2020- | .................... | .................... | .................... | .................... | .................... | .................... |
| M | 2050 hCoV-19/Austria/CeMM0179/2020\|EPI_ISL_583565\|2020- | .................... | .................... | .................... | .................... | .................... | .................... |
| M | 2051 hCoV-19/Austria/CeMM0316/2020\|EPI_ISL_583569\|2020- | .................... | .................... | .................... | .................... | .................... | .................... |
| M | 2052 hCoV-19/Austria/CeMM0488/2020\|EPI_ISL_583577\|2020- | .................... | .................... | .................... | .................... | .................... | .................... |
| M | 2053 hCoV-19/Austria/CeMM0489/2020\|EPI_ISL_583578\|2020- | .................... | .................... | .................... | .................... | .................... | .................... |
| M | 2054 hCoV-19/Austria/CeMM0490/2020\|EPI_ISL_583579\|2020- | .................... | .................... | .................... | .................... | .................... | .................... |
| M | 2055 hCoV-19/Austria/CeMM0491/2020\|EPI_ISL_583580\|2020- | .................... | .................... | .................... | .................... | .................... | .................... |
| M | 2056 hCoV-19/Austria/CeMM0492/2020\|EPI_ISL_583581\|2020- | .................... | .................... | .................... | .................... | .................... | .................... |
| M | 2057 hCoV-19/Austria/CeMM0493/2020\|EPI_ISL_583582\|2020- | .................... | .................... | .................... | .................... | .................... | .................... |
| M | 2058 hCoV-19/Austria/CeMM0494/2020\|EPI_ISL_583583\|2020- | .................... | .................... | .................... | .................... | .................... | .................... |
| M | 2059 hCoV-19/Austria/CeMM0495/2020\|EPI_ISL_583584\|2020- | .................... | .................... | .................... | .................... | .................... | .................... |
| M | 2060 hCoV-19/Austria/CeMM0496/2020\|EPI_ISL_583585\|2020- | .................... | .................... | .................... | .................... | .................... | .................... |
| M | 2061 hCoV-19/Austria/CeMM0497/2020\|EPI_ISL_583586\|2020- | .................... | .................... | .................... | .................... | .................... | .................... |
| M | 2062 hCoV-19/Austria/CeMM0499/2020\|EPI_ISL_583587\|2020- | .................... | .................... | .................... | .................... | .................... | .................... |
| M | 2063 hCoV-19/Austria/CeMM0500/2020\|EPI_ISL_583588\|2020- | .................... | .................... | .................... | .................... | .................... | .................... |
| M | 2064 hCoV-19/Austria/CeMM0501/2020\|EPI_ISL_583589\|2020- | .................... | .................... | .................... | .................... | .................... | .................... |
| M | 2065 hCoV-19/Austria/CeMM0503/2020\|EPI_ISL_583590\|2020- | .................... | .................... | .................... | .................... | .................... | .................... |
| M | 2066 hCoV-19/Austria/CeMM0504/2020\|EPI_ISL_583591\|2020- | .................... | .................... | .................... | .................... | .................... | .................... |
| M | 2067 hCoV-19/Austria/CeMM0505/2020\|EPI_ISL_583592\|2020- | .................... | .................... | .................... | .................... | .................... | .................... |
| M | 2068 hCoV-19/Austria/CeMM0506/2020\|EPI_ISL_583593\|2020- | .................... | .................... | .................... | .................... | .................... | .................... |
| M | 2069 hCoV-19/Austria/CeMM0508/2020\|EPI_ISL_583594\|2020- | .................... | .................... | .................... | .................... | .................... | .................... |
| M | 2070 hCoV-19/Austria/CeMM0509/2020\|EPI_ISL_583595\|2020- | .................... | .................... | .................... | .................... | .................... | .................... |
| M | 2071 hCoV-19/Austria/CeMM0510/2020\|EPI_ISL_583596\|2020- | .................... | .................... | .................... | .................... | .................... | .................... |
| M | 2072 hCoV-19/Austria/CeMM0512/2020\|EPI_ISL_583597\|2020- | .................... | .................... | .................... | .................... | .................... | .................... |
| M | 2073 hCoV-19/Austria/CeMM0513/2020\|EPI_ISL_583598\|2020- | .................... | .................... | .................... | .................... | .................... | .................... |
| M | 2074 hCoV-19/England/NORT-2A595B/2020\|EPI_ISL_584701\|20 | .................... | .A.................. | .................... | .................... | .................... | .................... |
| M | 2075 hCoV-19/England/NORT-2A59D3/2020\|EPI_ISL_584702\|20 | .................... | .................... | .................... | .................... | .................... | .................... |
| M | 2076 hCoV-19/England/NORT-2A5A85/2020\|EPI_ISL_584704\|20 | .................... | .................... | .................... | .................... | .................... | .................... |
| M | 2077 hCoV-19/England/NORT-2A5AB2/2020\|EPI_ISL_584705\|20 | .................... | .................... | .................... | .................... | .................... | .................... |
| M | 2078 hCoV-19/England/NORT-2A5C70/2020\|EPI_ISL_584707\|20 | .................... | .................... | .................... | .................... | .................... | .................... |
| M | 2079 hCoV-19/England/NORT-2A7C8D/2020\|EPI_ISL_584713\|20 | .................... | .................... | .................... | .................... | .................... | .................... |
| M | 2082 hCoV-19/England/NORW-F06E3/2020\|EPI_ISL_584737\|202 | .................... | .................... | .................... | .................... | .................... | .................... |
| M | 2085 hCoV-19/England/NOTT-11479A/2020\|EPI_ISL_584750\|20 | .................... | .................... | .................... | .................... | .................... | .................... |
| M | 2086 hCoV-19/England/NOTT-1147A9/2020\|EPI_ISL_584751\|20 | .................... | .................... | .................... | .................... | .................... | .................... |
| M | 2087 hCoV-19/England/NOTT-1147C7/2020\|EPI_ISL_584753\|20 | .................... | .................... | .................... | .................... | .................... | .................... |
| M | 2088 hCoV-19/England/NOTT-1148C4/2020\|EPI_ISL_584754\|20 | .................... | .................... | .................... | .................... | .................... | .................... |
| M | 2089 hCoV-19/England/NOTT-1148D3/2020\|EPI_ISL_584755\|20 | .................... | .................... | .................... | .................... | .................... | .................... |
| M | 2090 hCoV-19/England/NOTT-11493A/2020\|EPI_ISL_584756\|20 | .................... | .................... | .................... | .................... | .................... | .................... |

| | | | | | | | |
|---|---|---|---|---|---|---|---|
| M | 2091 hCoV-19/England/NOTT-114976/2020\|EPI_ISL_584757\|20 | .................... | .................... | ..................... | .................... | .................... | .................... |
| M | 2092 hCoV-19/England/NOTT-1149A3/2020\|EPI_ISL_584758\|20 | .................... | .................... | ..................... | .................... | .................... | .................... |
| M | 2093 hCoV-19/England/NOTT-1149C1/2020\|EPI_ISL_584759\|20 | .................... | .................... | ..................... | .................... | .................... | .................... |
| M | 2094 hCoV-19/England/NOTT-114A73/2020\|EPI_ISL_584760\|20 | .................... | .................... | ..................... | .................... | .................... | .................... |
| M | 2095 hCoV-19/England/NOTT-114ACE/2020\|EPI_ISL_584761\|20 | .................... | .................... | ..................... | .................... | .................... | .................... |
| M | 2096 hCoV-19/England/QEUH-A00507/2020\|EPI_ISL_584884\|20 | .................... | .................... | ..................... | .................... | .................... | .................... |
| M | 2097 hCoV-19/England/QEUH-A00561/2020\|EPI_ISL_584888\|20 | .................... | .................... | ..................... | .................... | .................... | .................... |
| M | 2098 hCoV-19/England/QEUH-A005AD/2020\|EPI_ISL_584889\|20 | .................... | .................... | ..................... | .................... | .................... | .................... |
| M | 2099 hCoV-19/England/QEUH-A005DA/2020\|EPI_ISL_584891\|20 | .................... | .................... | ..................... | .................... | .................... | .................... |
| M | 2100 hCoV-19/England/QEUH-A005F8/2020\|EPI_ISL_584892\|20 | .................... | .................... | ..................... | .................... | .................... | .................... |
| M | 2102 hCoV-19/England/QEUH-A00B41/2020\|EPI_ISL_584899\|20 | .................... | .................... | ..................... | .................... | .................... | .................... |
| M | 2103 hCoV-19/England/ALDP-A2FAB8/2020\|EPI_ISL_587112\|20 | .................... | .................... | ..................... | .................... | .................... | .................... |
| M | 2104 hCoV-19/England/ALDP-A2FB4C/2020\|EPI_ISL_587113\|20 | .................... | .................... | ..................... | .................... | .................... | .................... |
| M | 2105 hCoV-19/England/ALDP-A2ED74/2020\|EPI_ISL_587114\|20 | .................... | .A.................. | ..................... | .................... | .................... | ..A................. |
| M | 2106 hCoV-19/England/ALDP-A2EE53/2020\|EPI_ISL_587115\|20 | .................... | .................... | ..................... | .................... | .................... | .................... |
| M | 2107 hCoV-19/England/QEUH-9FABC3/2020\|EPI_ISL_588227\|20 | .................... | .................... | ..................... | .................... | .................... | .................... |
| M | 2108 hCoV-19/England/QEUH-9FAFF4/2020\|EPI_ISL_588229\|20 | .................... | .................... | ..................... | .................... | .................... | .................... |
| M | 2109 hCoV-19/England/QEUH-9FB11B/2020\|EPI_ISL_588230\|20 | .................... | .................... | ..................... | .................... | .................... | .................... |
| M | 2110 hCoV-19/England/QEUH-9FAC48/2020\|EPI_ISL_588231\|20 | .................... | .................... | ..................... | .................... | .................... | .................... |
| M | 2111 hCoV-19/England/QEUH-9FAF5E/2020\|EPI_ISL_588232\|20 | .................... | .................... | ..................... | .................... | .................... | .................... |
| M | 2112 hCoV-19/England/QEUH-9FA60E/2020\|EPI_ISL_588233\|20 | .................... | .................... | ..................... | .................... | .................... | .................... |
| M | 2113 hCoV-19/England/QEUH-9FAAD5/2020\|EPI_ISL_588234\|20 | .................... | .................... | ..................... | .................... | ......T............. | .................... |
| M | 2114 hCoV-19/England/QEUH-9FB12A/2020\|EPI_ISL_588235\|20 | .................... | .................... | ..................... | .................... | .................... | .................... |
| M | 2115 hCoV-19/England/QEUH-9FB4C7/2020\|EPI_ISL_588237\|20 | .................... | .................... | ..................... | .................... | .................... | .................... |
| M | 2116 hCoV-19/England/QEUH-9F4956/2020\|EPI_ISL_588290\|20 | .................... | .................... | ..................... | .................... | .................... | .................... |
| M | 2117 hCoV-19/England/QEUH-9F4CF3/2020\|EPI_ISL_588291\|20 | .................... | .................... | ..................... | .................... | .................... | .................... |
| M | 2118 hCoV-19/England/QEUH-9F5BAA/2020\|EPI_ISL_588292\|20 | .................... | .................... | ..................... | .................... | .................... | .................... |
| M | 2119 hCoV-19/England/QEUH-9F48D1/2020\|EPI_ISL_588293\|20 | .................... | .................... | ..................... | .................... | .................... | .................... |
| M | 2120 hCoV-19/England/QEUH-9F5BB9/2020\|EPI_ISL_588294\|20 | .................... | .A.................. | ..................... | .................... | .................... | ..A................. |
| M | 2121 hCoV-19/England/QEUH-9F4FEB/2020\|EPI_ISL_588295\|20 | .................... | .................... | ..................... | .................... | .................... | .................... |
| M | 2122 hCoV-19/England/QEUH-9F48E0/2020\|EPI_ISL_588296\|20 | .................... | .................... | ..................... | .................... | .................... | .................... |
| M | 2123 hCoV-19/England/QEUH-9F4974/2020\|EPI_ISL_588297\|20 | .................... | .A.................. | ..................... | .................... | .................... | .................... |
| M | 2124 hCoV-19/England/QEUH-9F5E83/2020\|EPI_ISL_588299\|20 | .................... | .................... | ..................... | .................... | .................... | .................... |
| M | 2125 hCoV-19/England/QEUH-9F491A/2020\|EPI_ISL_588403\|20 | .................... | .................... | ..................... | .................... | ....T............... | .................... |
| M | 2126 hCoV-19/England/QEUH-9F5B9B/2020\|EPI_ISL_588404\|20 | .................... | .................... | ..................... | .................... | ......T............. | .................... |
| M | 2127 hCoV-19/England/QEUH-9F483B/2020\|EPI_ISL_588405\|20 | .................... | .................... | ..................... | .................... | .................... | .................... |
| M | 2128 hCoV-19/England/QEUH-9F4EDF/2020\|EPI_ISL_588406\|20 | .................... | .................... | ..................... | .................... | .................... | .................... |
| M | 2129 hCoV-19/England/QEUH-9F4A80/2020\|EPI_ISL_588407\|20 | .................... | .................... | ..................... | .................... | .................... | .................... |
| M | 2130 hCoV-19/England/QEUH-9F4ABD/2020\|EPI_ISL_588408\|20 | .................... | .A.................. | ..................... | .................... | .................... | .................... |
| M | 2131 hCoV-19/England/QEUH-9F45AD/2020\|EPI_ISL_588409\|20 | .................... | .................... | ..................... | .................... | .................... | .................... |
| M | 2132 hCoV-19/England/QEUH-9F45CB/2020\|EPI_ISL_588410\|20 | .................... | .................... | ..................... | .................... | .................... | .................... |
| M | 2133 hCoV-19/England/QEUH-9F4D1E/2020\|EPI_ISL_588411\|20 | .................... | .................... | ..................... | .................... | ......T............. | .................... |
| M | 2134 hCoV-19/England/QEUH-9F5E74/2020\|EPI_ISL_588412\|20 | .................... | .................... | ..................... | .................... | .................... | .................... |
| M | 2135 hCoV-19/England/QEUH-9F49ED/2020\|EPI_ISL_588413\|20 | .................... | .A.................. | ..................... | .................... | .................... | .................... |
| M | 2136 hCoV-19/England/QEUH-9F4B9C/2020\|EPI_ISL_588414\|20 | .................... | .................... | ..................... | .................... | .................... | .................... |
| M | 2137 hCoV-19/England/QEUH-9F47C5/2020\|EPI_ISL_588415\|20 | .................... | .................... | ..................... | .................... | .................... | .................... |
| M | 2138 hCoV-19/England/QEUH-9F465F/2020\|EPI_ISL_588416\|20 | .................... | .................... | ..................... | .................... | .................... | .................... |
| M | 2139 hCoV-19/England/QEUH-9F5C4D/2020\|EPI_ISL_588417\|20 | .................... | .................... | ..................... | .................... | .................... | .................... |
| M | 2140 hCoV-19/England/QEUH-9F5C10/2020\|EPI_ISL_588418\|20 | .................... | .................... | ..................... | .................... | .................... | .................... |
| M | 2141 hCoV-19/England/QEUH-9F47E3/2020\|EPI_ISL_588420\|20 | .................... | .................... | ..................... | .................... | .................... | .................... |
| M | 2142 hCoV-19/England/QEUH-9F666C/2020\|EPI_ISL_588421\|20 | .................... | .................... | ..................... | .................... | .................... | .................... |
| M | 2143 hCoV-19/England/QEUH-9F583A/2020\|EPI_ISL_588422\|20 | .................... | .................... | ..................... | .................... | .................... | .................... |
| M | 2144 hCoV-19/England/QEUH-9F58D0/2020\|EPI_ISL_588423\|20 | .................... | .................... | ..................... | .................... | .................... | .................... |
| M | 2145 hCoV-19/England/QEUH-9F64AE/2020\|EPI_ISL_588440\|20 | .................... | .................... | ..................... | .................... | .................... | .................... |
| M | 2146 hCoV-19/England/QEUH-9F44A0/2020\|EPI_ISL_588443\|20 | .................... | .................... | ..................... | .................... | .................... | .................... |
| M | 2147 hCoV-19/England/QEUH-9F5278/2020\|EPI_ISL_588444\|20 | .................... | .................... | ..................... | .................... | ....T............... | .................... |
| M | 2148 hCoV-19/Wales/QEUH-9F6480/2020\|EPI_ISL_588445\|2020 | .................... | .................... | ..................... | .................... | .................... | .................... |
| M | 2149 hCoV-19/England/QEUH-9F6505/2020\|EPI_ISL_588446\|20 | .................... | .................... | ..................... | .................... | .................... | .................... |
| M | 2150 hCoV-19/England/ALDP-9EE0E1/2020\|EPI_ISL_589558\|20 | .................... | .................... | ..................... | .................... | .................... | .................... |
| M | 2151 hCoV-19/England/ALDP-9ED361/2020\|EPI_ISL_589560\|20 | ...T................ | .................... | ..................... | .................... | .................... | .................... |
| M | 2152 hCoV-19/England/ALDP-9ECF04/2020\|EPI_ISL_589561\|20 | .................... | .................... | ..................... | .................... | .................... | .................... |
| M | 2153 hCoV-19/England/ALDP-9ED4AA/2020\|EPI_ISL_589562\|20 | .................... | .................... | ..................... | .................... | .................... | .................... |
| M | 2154 hCoV-19/England/ALDP-9EE3D9/2020\|EPI_ISL_589563\|20 | .................... | .................... | ..................... | .................... | .................... | .................... |
| M | 2155 hCoV-19/England/ALDP-9EE10C/2020\|EPI_ISL_589565\|20 | .................... | .................... | ..................... | .................... | .................... | .................... |
| M | 2156 hCoV-19/England/ALDP-9EDEAC/2020\|EPI_ISL_589568\|20 | .................... | .................... | ..................... | .................... | .................... | .................... |
| M | 2157 hCoV-19/England/MILK-9F1151/2020\|EPI_ISL_589734\|20 | .................... | .................... | ..................... | .................... | .................... | .................... |
| M | 2158 hCoV-19/England/MILK-9EF569/2020\|EPI_ISL_589735\|20 | .................... | .................... | ..................... | .................... | .................... | .................... |
| M | 2159 hCoV-19/England/MILK-9F0C06/2020\|EPI_ISL_589736\|20 | .................... | .................... | ..................... | .................... | .................... | .................... |
| M | 2160 hCoV-19/England/MILK-9F1F0C/2020\|EPI_ISL_589738\|20 | .................... | .................... | ..................... | .................... | .................... | .................... |
| M | 2161 hCoV-19/England/MILK-9F134C/2020\|EPI_ISL_589739\|20 | .................... | .................... | ..................... | .................... | .................... | .................... |
| M | 2162 hCoV-19/England/MILK-9F1B26/2020\|EPI_ISL_589740\|20 | .................... | .................... | ..................... | .................... | .................... | .................... |
| M | 2163 hCoV-19/England/MILK-9F095A/2020\|EPI_ISL_589741\|20 | .................... | .................... | ..................... | .................... | .................... | .................... |
| M | 2164 hCoV-19/England/MILK-9F191D/2020\|EPI_ISL_589742\|20 | .................... | .A.................. | ..................... | .................... | .................... | ..A................. |
| M | 2165 hCoV-19/England/MILK-9F168F/2020\|EPI_ISL_589743\|20 | .................... | .................... | ..................... | .................... | .................... | .................... |
| M | 2166 hCoV-19/England/MILK-9F0CCA/2020\|EPI_ISL_589744\|20 | .................... | .................... | ..................... | .................... | .................... | .................... |
| M | 2167 hCoV-19/England/MILK-9F1027/2020\|EPI_ISL_589745\|20 | .................... | .................... | ..................... | .................... | .................... | .................... |
| M | 2168 hCoV-19/England/MILK-9F1625/2020\|EPI_ISL_589746\|20 | .................... | .................... | ..................... | .................... | .................... | .................... |
| M | 2169 hCoV-19/England/MILK-9F1D8A/2020\|EPI_ISL_589747\|20 | .................... | .................... | ..................... | .................... | .................... | .................... |
| M | 2170 hCoV-19/England/MILK-9F10FA/2020\|EPI_ISL_589748\|20 | .................... | .................... | ..................... | .................... | .................... | .................... |
| M | 2171 hCoV-19/England/MILK-9F092D/2020\|EPI_ISL_589749\|20 | .................... | .................... | ..................... | .................... | .................... | .................... |

```
M    2172 hCoV-19/England/MILK-9F1BBD/2020|EPI_ISL_589750|20   ····················  ····················  ····················  ····················  ····················  ····················
M    2173 hCoV-19/England/MILK-9F0AEE/2020|EPI_ISL_589751|20   ····················  ····················  ····················  ····················  ····················  ····················
M    2174 hCoV-19/England/MILK-9F0BBE/2020|EPI_ISL_589752|20   ····················  ····················  ····················  ····················  ·····T··············  ····················
M    2175 hCoV-19/England/MILK-9F0B54/2020|EPI_ISL_589753|20   ····················  ····················  ····················  ····················  ····················  ····················
M    2176 hCoV-19/England/MILK-9F0F2B/2020|EPI_ISL_589754|20   ····················  ·A··················  ····················  ····················  ····················  ··A·················
M    2177 hCoV-19/England/MILK-9F1634/2020|EPI_ISL_589755|20   ····················  ····················  ····················  ····················  ····················  ····················
M    2178 hCoV-19/England/QEUH-9E40AF/2020|EPI_ISL_589774|20   ····················  ····················  ····················  ····················  ···T················  ····················
M    2179 hCoV-19/England/QEUH-9E53E1/2020|EPI_ISL_589775|20   ····················  ····················  ····················  ····················  ····················  ····················
M    2180 hCoV-19/England/QEUH-9E53A5/2020|EPI_ISL_589776|20   ····················  ····················  ····················  ····················  ····················  ····················
M    2181 hCoV-19/England/QEUH-9E492C/2020|EPI_ISL_589777|20   ····················  ····················  ····················  ····················  ····················  ····················
M    2182 hCoV-19/England/QEUH-9B7746/2020|EPI_ISL_590540|20   ····················  ····················  ····················  ····················  ·····T··············  ····················
M    2183 hCoV-19/England/QEUH-9B749A/2020|EPI_ISL_590541|20   ····················  ····················  ····················  ····················  ····················  ····················
M    2184 hCoV-19/England/QEUH-9B7597/2020|EPI_ISL_590542|20   ····················  ····················  ····················  ····················  ····················  ····················
M    2185 hCoV-19/England/QEUH-9B7BD1/2020|EPI_ISL_590543|20   ····················  ····················  ····················  ····················  ····················  ····················
M    2186 hCoV-19/England/QEUH-9B7B68/2020|EPI_ISL_590544|20   ····················  ····················  ····················  ····················  ····················  ····················
M    2187 hCoV-19/England/QEUH-9B763A/2020|EPI_ISL_590546|20   ····················  ····················  ····················  ····················  ····················  ····················
M    2188 hCoV-19/England/QEUH-9B6FC7/2020|EPI_ISL_590549|20   ····················  ····················  ····················  ····················  ····················  ····················
M    2189 hCoV-19/USA/MI-UM-10036245036/2020|EPI_ISL_590721|   ····················  ····················  ····················  ····················  ····················  ····················
M    2191 hCoV-19/USA/MI-UM-10036226294/2020|EPI_ISL_590723|   ····················  ·A··················  ····················  ····················  ····················  ····················
M    2192 hCoV-19/USA/MI-UM-10036329024/2020|EPI_ISL_590724|   ····················  ····················  ····················  ····················  ····················  ····················
M    2193 hCoV-19/USA/MI-UM-10036150967/2020|EPI_ISL_590726|   ····················  ····················  ····················  ····················  ····················  ····················
M    2194 hCoV-19/USA/MI-UM-10036155827/2020|EPI_ISL_590727|   ····················  ·············A······  ····················  ····················  ····················  ····················
M    2195 hCoV-19/USA/MI-UM-10036150256/2020|EPI_ISL_590728|   ····················  ····················  ····················  ····················  ····················  ····················
M    2197 hCoV-19/USA/MI-UM-10036173099/2020|EPI_ISL_590730|   ····················  ····················  ····················  ····················  ····················  ····················
M    2198 hCoV-19/USA/MI-UM-10036168091/2020|EPI_ISL_590731|   ····················  ····················  ····················  ····················  ····················  ····················
M    2199 hCoV-19/USA/MI-UM-10036150579/2020|EPI_ISL_590732|   ····················  ····················  ····················  ····················  ····················  ····················
M    2200 hCoV-19/USA/MI-UM-10036165196/2020|EPI_ISL_590734|   ····················  ····················  ····················  ····················  ····················  ····················
M    2202 hCoV-19/USA/MI-UM-10036206650/2020|EPI_ISL_590736|   ····················  ·A··················  ····················  ····················  ····················  ····················
M    2203 hCoV-19/USA/MI-UM-10036173189/2020|EPI_ISL_590737|   ····················  ·A··················  ····················  ····················  ····················  ····················
M    2204 hCoV-19/USA/MI-UM-10036205311/2020|EPI_ISL_590738|   ····················  ····················  ····················  ····················  ····················  ····················
M    2205 hCoV-19/USA/MI-UM-10036195123/2020|EPI_ISL_590739|   ····················  ·············A······  ····················  ····················  ····················  ····················
M    2207 hCoV-19/USA/MN-MDH-1749/2020|EPI_ISL_590754|2020-0   ····················  ····················  ····················  ····················  ····················  ····················
M    2208 hCoV-19/USA/MN-MDH-1750/2020|EPI_ISL_590755|2020-0   ····················  ····················  ····················  ····················  ····················  ····················
M    2209 hCoV-19/USA/MN-MDH-1751/2020|EPI_ISL_590756|2020-0   ····················  ····················  ····················  ····················  ·NN·················  ····················
M    2210 hCoV-19/USA/MN-MDH-1752/2020|EPI_ISL_590757|2020-0   ····················  ····················  ····················  ····················  ····················  ····················
M    2211 hCoV-19/USA/MN-MDH-1753/2020|EPI_ISL_590758|2020-0   ····················  ····················  ····················  ····················  ····················  ····················
M    2212 hCoV-19/USA/MN-MDH-1754/2020|EPI_ISL_590759|2020-0   ····················  ····················  ····················  ····················  ····················  ····················
M    2213 hCoV-19/Australia/VIC11833/2020|EPI_ISL_591870|202   ····················  ····················  ····················  ····················  ····················  ····················
M    2214 hCoV-19/Australia/VIC11835/2020|EPI_ISL_591871|202   ····················  ····················  ····················  ····················  ····················  ····················
M    2215 hCoV-19/Australia/VIC11841/2020|EPI_ISL_591872|202   ····················  ····················  ····················  ····················  ····················  ····················
M    2216 hCoV-19/Australia/VIC11854/2020|EPI_ISL_591873|202   ····················  ····················  ····················  ····················  ····················  ····················
M    2217 hCoV-19/Australia/VIC11856/2020|EPI_ISL_591874|202   ····················  ····················  ···K················  ····················  ····················  ····················
M    2218 hCoV-19/Australia/VIC11857/2020|EPI_ISL_591875|202   ····················  ····················  ···K················  ····················  ····················  ····················
M    2219 hCoV-19/Australia/VIC11868/2020|EPI_ISL_591877|202   ····················  ····················  ···K················  ····················  ····················  ····················
M    2220 hCoV-19/USA/UT-UPHL-201024487/2020|EPI_ISL_594248|   ····················  ····················  ····················  ····················  ····················  ····················
M    2221 hCoV-19/USA/UT-UPHL-201028884/2020|EPI_ISL_594252|   ····················  ····················  ····················  ····················  ········A···········  ····················
M    2222 hCoV-19/USA/UT-UPHL-201022611/2020|EPI_ISL_594253|   ····················  ····················  ····················  ····················  ········A···········  ····················
M    2223 hCoV-19/USA/UT-UPHL-201025480/2020|EPI_ISL_594255|   ····················  ····················  ····················  ····················  ····················  ····················
M    2224 hCoV-19/USA/UT-UPHL-201016621/2020|EPI_ISL_594256|   ····················  ·A··················  ····················  ····················  ····················  ····················
M    2225 hCoV-19/USA/UT-UPHL-201014927/2020|EPI_ISL_594258|   ····················  ····················  ····················  ····················  ····················  ····················
M    2226 hCoV-19/USA/UT-UPHL-201014403/2020|EPI_ISL_594259|   ····················  ····················  ····················  ····················  ····················  ····················
M    2227 hCoV-19/USA/UT-UPHL-201012311/2020|EPI_ISL_594260|   ····················  ····················  ····················  ····················  ····················  ····················
M    2228 hCoV-19/USA/UT-UPHL-201025399/2020|EPI_ISL_594263|   ····················  ····················  ····················  ····················  ····················  ····················
M    2229 hCoV-19/USA/UT-UPHL-201019147/2020|EPI_ISL_594266|   ····················  ·A··················  ····················  ····················  ····················  ····················
M    2230 hCoV-19/USA/UT-UPHL-201004526/2020|EPI_ISL_594267|   ····················  ·A··················  ····················  ····················  ····················  ····················
M    2231 hCoV-19/USA/UT-UPHL-201030232/2020|EPI_ISL_594268|   ····················  ····················  ····················  ····················  ····················  ····················
M    2232 hCoV-19/USA/UT-UPHL-201021766/2020|EPI_ISL_594270|   ····················  ····················  ····················  ····················  ····················  ····················
M    2233 hCoV-19/USA/UT-UPHL-201026014/2020|EPI_ISL_594271|   ····················  ····················  ····················  ····················  ····················  ····················
M    2234 hCoV-19/USA/UT-UPHL-201021139/2020|EPI_ISL_594273|   ····················  ·A··················  ····················  ····················  ····················  ····················
M    2235 hCoV-19/USA/UT-UPHL-201007266/2020|EPI_ISL_594274|   ····················  ····················  ····················  ····················  ····················  ····················
M    2236 hCoV-19/USA/UT-UPHL-201002531/2020|EPI_ISL_594275|   ····················  ····················  ····················  ····················  ····················  ····················
M    2237 hCoV-19/USA/FL-BPHL-1589/2020|EPI_ISL_594279|2020-   ····················  ····················  ····················  ····················  ····················  ····················
M    2238 hCoV-19/USA/FL-BPHL-1590/2020|EPI_ISL_594280|2020-   ····················  ····················  ····················  ····················  ····················  ····················
M    2239 hCoV-19/USA/FL-BPHL-1591/2020|EPI_ISL_594281|2020-   ····················  ····················  ····················  ····················  ····················  ····················
M    2240 hCoV-19/USA/FL-BPHL-1592/2020|EPI_ISL_594282|2020-   ····················  ····················  ····················  ····················  ····················  ····················
M    2241 hCoV-19/USA/FL-BPHL-1594/2020|EPI_ISL_594284|2020-   ····················  ····················  ····················  ····················  ····················  ····················
M    2242 hCoV-19/Wales/PHWC-47E081/2020|EPI_ISL_595400|2020   ····················  ····················  ····················  ····················  ····················  ····················
M    2243 hCoV-19/Wales/PHWC-47E090/2020|EPI_ISL_595401|2020   ····················  ····················  ····················  ····················  ····················  ····················
M    2244 hCoV-19/Wales/PHWC-47E0AF/2020|EPI_ISL_595402|2020   ····················  ····················  ····················  ····················  ····················  ····················
M    2245 hCoV-19/Wales/PHWC-47E0BE/2020|EPI_ISL_595403|2020   ····················  ····················  ····················  ····················  ····················  ····················
M    2246 hCoV-19/Wales/PHWC-47E0CD/2020|EPI_ISL_595404|2020   ····················  ····················  ····················  ····················  ····················  ····················
M    2247 hCoV-19/Wales/PHWC-47E0DC/2020|EPI_ISL_595405|2020   ····················  ····················  ····················  ····················  ····················  ····················
M    2248 hCoV-19/Wales/PHWC-47E0EB/2020|EPI_ISL_595406|2020   ····················  ····················  ····················  ····················  ····················  ····················
M    2249 hCoV-19/Wales/PHWC-47E0FA/2020|EPI_ISL_595407|2020   ····················  ····················  ····················  ····················  ····················  ····················
M    2250 hCoV-19/Wales/PHWC-47E106/2020|EPI_ISL_595408|2020   ····················  ····················  ····················  ····················  ····················  ····················
M    2251 hCoV-19/Wales/PHWC-47E115/2020|EPI_ISL_595409|2020   ····················  ····················  ····················  ····················  ····················  ····················
M    2252 hCoV-19/Wales/PHWC-47E9B3/2020|EPI_ISL_595532|2020   ····················  ····················  ····················  ····················  ····················  ····················
M    2254 hCoV-19/Wales/PHWC-47E9D1/2020|EPI_ISL_595534|2020   ····················  ····················  ····················  ····················  ····················  ····················
M    2255 hCoV-19/Wales/PHWC-47E9FF/2020|EPI_ISL_595536|2020   ····················  ····················  ····················  ····················  ····················  ····················
M    2257 hCoV-19/Wales/PHWC-47EA1A/2020|EPI_ISL_595538|2020   ····················  ····················  ····················  ····················  ····················  ····················
```

| | | | | | | | |
|---|---|---|---|---|---|---|---|
| M | 2258 hCoV-19/Wales/PHWC-47EA47/2020\|EPI_ISL_595541\|2020 | ···················· | ···················· | ···················· | ···················· | ···················· | ···················· |
| M | 2259 hCoV-19/Wales/PHWC-47EA56/2020\|EPI_ISL_595542\|2020 | ···················· | ···················· | ···················· | ···················· | ···················· | ···················· |
| M | 2260 hCoV-19/Wales/PHWC-47EA65/2020\|EPI_ISL_595543\|2020 | ···················· | ···················· | ···················· | ···················· | ···················· | ···················· |
| M | 2261 hCoV-19/Wales/PHWC-47EA83/2020\|EPI_ISL_595545\|2020 | ···················· | ···················· | ···················· | ···················· | ···················· | ···················· |
| M | 2262 hCoV-19/Wales/PHWC-47EAA1/2020\|EPI_ISL_595547\|2020 | ···················· | ···················· | ···················· | ···················· | ···················· | ···················· |
| M | 2263 hCoV-19/Wales/PHWC-47EAB0/2020\|EPI_ISL_595548\|2020 | ···················· | ···················· | ···················· | ···················· | ···················· | ···················· |
| M | 2264 hCoV-19/Wales/PHWC-47EADE/2020\|EPI_ISL_595550\|2020 | ···················· | ···················· | ···················· | ···················· | ···················· | ···················· |
| M | 2265 hCoV-19/Wales/PHWC-47EAED/2020\|EPI_ISL_595551\|2020 | ···················· | ···················· | ···················· | ···················· | ···················· | ···················· |
| M | 2266 hCoV-19/Wales/PHWC-47EAFC/2020\|EPI_ISL_595552\|2020 | ···················· | ···················· | ···················· | ···················· | ···················· | ···················· |
| M | 2267 hCoV-19/Wales/PHWC-47EB08/2020\|EPI_ISL_595553\|2020 | ···················· | ···················· | ···················· | ···················· | ···················· | ···················· |
| M | 2268 hCoV-19/Wales/PHWC-47EB17/2020\|EPI_ISL_595554\|2020 | ···················· | ···················· | ···················· | ···················· | ···················· | ···················· |
| M | 2269 hCoV-19/Wales/PHWC-47EB26/2020\|EPI_ISL_595555\|2020 | ···················· | ···················· | ···················· | ···················· | ···················· | ···················· |
| M | 2270 hCoV-19/Wales/PHWC-47EB35/2020\|EPI_ISL_595556\|2020 | ···················· | ···················· | ···················· | ···················· | ···················· | ···················· |
| M | 2272 hCoV-19/Wales/PHWC-47EB62/2020\|EPI_ISL_595559\|2020 | ···················· | ···················· | ···················· | ···················· | ···················· | ···················· |
| M | 2273 hCoV-19/Wales/PHWC-47EB71/2020\|EPI_ISL_595560\|2020 | ···················· | ···················· | ···················· | ···················· | ···················· | ···················· |
| M | 2274 hCoV-19/Wales/PHWC-47EB80/2020\|EPI_ISL_595561\|2020 | ···················· | ···················· | ···················· | ···················· | ···················· | ···················· |
| M | 2275 hCoV-19/Wales/PHWC-47EB9F/2020\|EPI_ISL_595562\|2020 | ···················· | ···················· | ···················· | ···················· | ···················· | ···················· |
| M | 2276 hCoV-19/Wales/PHWC-47EBAE/2020\|EPI_ISL_595563\|2020 | ···················· | ···················· | ···················· | ···················· | ···················· | ···················· |
| M | 2277 hCoV-19/Wales/PHWC-47EBBD/2020\|EPI_ISL_595564\|2020 | ···················· | ···················· | ···················· | ···················· | ···················· | ···················· |
| M | 2278 hCoV-19/Wales/PHWC-47EBCC/2020\|EPI_ISL_595565\|2020 | ···················· | ···················· | ···················· | ···················· | ···················· | ···················· |
| M | 2279 hCoV-19/Wales/PHWC-47EBF9/2020\|EPI_ISL_595568\|2020 | ···················· | ···················· | ···················· | ···················· | ···················· | ···················· |
| M | 2280 hCoV-19/Wales/PHWC-47EC23/2020\|EPI_ISL_595570\|2020 | ···················· | ···················· | ···················· | ···················· | ···················· | ···················· |
| M | 2281 hCoV-19/Wales/PHWC-47EC32/2020\|EPI_ISL_595571\|2020 | ···················· | ···················· | ···················· | ···················· | ···················· | ···················· |
| M | 2282 hCoV-19/Wales/PHWC-47EC50/2020\|EPI_ISL_595572\|2020 | ···················· | ···················· | ···················· | ···················· | ···················· | ···················· |
| M | 2283 hCoV-19/Wales/PHWC-47EC7E/2020\|EPI_ISL_595573\|2020 | ···················· | ···················· | ···················· | ···················· | ···················· | ···················· |
| M | 2284 hCoV-19/Wales/PHWC-47ECAB/2020\|EPI_ISL_595576\|2020 | ···················· | ···················· | ···················· | ···················· | ···················· | ···················· |
| M | 2285 hCoV-19/Wales/PHWC-47ECC9/2020\|EPI_ISL_595577\|2020 | ···················· | ···················· | ···················· | ···················· | ···················· | ···················· |
| M | 2286 hCoV-19/Wales/PHWC-47ECD8/2020\|EPI_ISL_595578\|2020 | ···················· | ···················· | ···················· | ···················· | ···················· | ···················· |
| M | 2287 hCoV-19/Wales/PHWC-47ECE7/2020\|EPI_ISL_595579\|2020 | ···················· | ···················· | ···················· | ···················· | ···················· | ···················· |
| M | 2288 hCoV-19/Wales/PHWC-47ECF6/2020\|EPI_ISL_595580\|2020 | ···················· | ···················· | ···················· | ···················· | ···················· | ···················· |
| M | 2289 hCoV-19/Wales/PHWC-47ED20/2020\|EPI_ISL_595582\|2020 | ···················· | ···················· | ···················· | ···················· | ···················· | ···················· |
| M | 2291 hCoV-19/Wales/PHWC-47ED4E/2020\|EPI_ISL_595584\|2020 | ···················· | ···················· | ···················· | ···················· | ···················· | ···················· |
| M | 2293 hCoV-19/Wales/PHWC-47ED6C/2020\|EPI_ISL_595586\|2020 | ···················· | ···················· | ···················· | ···················· | ···················· | ···················· |
| M | 2294 hCoV-19/Wales/PHWC-47ED8A/2020\|EPI_ISL_595587\|2020 | ···················· | ···················· | ···················· | ···················· | ···················· | ···················· |
| M | 2295 hCoV-19/Wales/PHWC-47ED99/2020\|EPI_ISL_595588\|2020 | ···················· | ···················· | ···················· | ···················· | ···················· | ···················· |
| M | 2296 hCoV-19/Wales/PHWC-47EDA8/2020\|EPI_ISL_595589\|2020 | ···················· | ···················· | ···················· | ···················· | ···················· | ···················· |
| M | 2297 hCoV-19/Wales/PHWC-47EDB7/2020\|EPI_ISL_595590\|2020 | ···················· | ···················· | ···················· | ···················· | ···················· | ···················· |
| M | 2298 hCoV-19/Wales/PHWC-47EDC6/2020\|EPI_ISL_595591\|2020 | ···················· | ···················· | ···················· | ···················· | ···················· | ···················· |
| M | 2299 hCoV-19/Wales/PHWC-47EDD5/2020\|EPI_ISL_595592\|2020 | ···················· | ···················· | ···················· | ···················· | ···················· | ···················· |
| M | 2300 hCoV-19/Wales/PHWC-47EDF3/2020\|EPI_ISL_595594\|2020 | ···················· | ···················· | ···················· | ···················· | ···················· | ···················· |
| M | 2301 hCoV-19/Wales/PHWC-47EE0F/2020\|EPI_ISL_595595\|2020 | ···················· | ···················· | ···················· | ···················· | ···················· | ···················· |
| M | 2302 hCoV-19/Wales/PHWC-47EE1E/2020\|EPI_ISL_595596\|2020 | ···················· | ···················· | ···················· | ···················· | ···················· | ···················· |
| M | 2303 hCoV-19/Wales/PHWC-47EE3C/2020\|EPI_ISL_595598\|2020 | ···················· | ···················· | ···················· | ···················· | ···················· | ···················· |
| M | 2304 hCoV-19/Wales/PHWC-47EE4B/2020\|EPI_ISL_595599\|2020 | ···················· | ···················· | ···················· | ···················· | ···················· | ···················· |
| M | 2305 hCoV-19/England/QEUH-9FA756/2020\|EPI_ISL_588205\|20 | ···················· | ···················· | ···················· | ···················· | ···················· | ···················· |
| M | 2306 hCoV-19/England/QEUH-9FAF03/2020\|EPI_ISL_588206\|20 | ···················· | ···················· | ···················· | ···················· | ···················· | ···················· |
| M | 2307 hCoV-19/England/QEUH-9FAEE8/2020\|EPI_ISL_588208\|20 | ···················· | ···················· | ···················· | ···················· | ···················· | ···················· |
| M | 2308 hCoV-19/England/QEUH-9FA48C/2020\|EPI_ISL_588209\|20 | ···················· | ·A·················· | ···················· | ···················· | ···················· | ··A················· |
| M | 2309 hCoV-19/England/QEUH-9FAE42/2020\|EPI_ISL_588210\|20 | ···················· | ···················· | ···················· | ···················· | ···················· | ···················· |
| M | 2310 hCoV-19/England/QEUH-9FB0A5/2020\|EPI_ISL_588212\|20 | ···················· | ···················· | ···················· | ···················· | ···················· | ···················· |
| M | 2311 hCoV-19/England/QEUH-9FA738/2020\|EPI_ISL_588213\|20 | ···················· | ···················· | ···················· | ···················· | ···················· | ···················· |
| M | 2312 hCoV-19/England/QEUH-9FB4B8/2020\|EPI_ISL_588214\|20 | ···················· | ···················· | ···················· | ···················· | ···················· | ···················· |
| M | 2313 hCoV-19/England/QEUH-9FAEAC/2020\|EPI_ISL_588215\|20 | ···················· | ···················· | ···················· | ···················· | ···················· | ···················· |
| M | 2314 hCoV-19/England/QEUH-9FB078/2020\|EPI_ISL_588216\|20 | ···················· | ···················· | ···················· | ···················· | ···················· | ···················· |
| M | 2315 hCoV-19/England/QEUH-9FA370/2020\|EPI_ISL_588217\|20 | ···················· | ···················· | ···················· | ···················· | ···················· | ···················· |
| M | 2316 hCoV-19/England/QEUH-9FAE24/2020\|EPI_ISL_588218\|20 | ···················· | ···················· | ···················· | ···················· | ···················· | ···················· |
| M | 2317 hCoV-19/England/QEUH-9FABB4/2020\|EPI_ISL_588219\|20 | ···················· | ···················· | ···················· | ···················· | ···················· | ···················· |
| M | 2318 hCoV-19/England/QEUH-9FA7A1/2020\|EPI_ISL_588220\|20 | ···················· | ···················· | ···················· | ···················· | ···················· | ···················· |
| M | 2319 hCoV-19/England/QEUH-9FB0E1/2020\|EPI_ISL_588221\|20 | ···················· | ···················· | ···················· | ···················· | ···················· | ···················· |
| M | 2320 hCoV-19/England/QEUH-9FB04B/2020\|EPI_ISL_588222\|20 | ···················· | ···················· | ···················· | ···················· | ···················· | ···················· |
| M | 2321 hCoV-19/England/QEUH-9FB4F4/2020\|EPI_ISL_588223\|20 | ···················· | ···················· | ···················· | ···················· | ·····T·············· | ···················· |
| M | 2322 hCoV-19/England/QEUH-9FAB5A/2020\|EPI_ISL_588224\|20 | ···················· | ···················· | ···················· | ···················· | ···················· | ···················· |
| M | 2323 hCoV-19/England/QEUH-9FABE1/2020\|EPI_ISL_588225\|20 | ···················· | ···················· | ···················· | ···················· | ···················· | ···················· |
| M | 2324 hCoV-19/England/QEUH-9FAE7F/2020\|EPI_ISL_588226\|20 | ···················· | ···················· | ···················· | ···················· | ···················· | ···················· |
| M | 2325 hCoV-19/England/QEUH-9FA5A7/2020\|EPI_ISL_588238\|20 | ···················· | ···················· | ···················· | ···················· | ···················· | ···················· |
| M | 2326 hCoV-19/England/QEUH-9FA307/2020\|EPI_ISL_588239\|20 | ···················· | ···················· | ···················· | ···················· | ···················· | ···················· |
| M | 2327 hCoV-19/England/QEUH-9FAF6D/2020\|EPI_ISL_588240\|20 | ···················· | ···················· | ···················· | ···················· | ···················· | ···················· |
| M | 2328 hCoV-19/England/QEUH-9FA334/2020\|EPI_ISL_588241\|20 | ···················· | ···················· | ···················· | ···················· | ···················· | ···················· |
| M | 2329 hCoV-19/England/QEUH-9FAD63/2020\|EPI_ISL_588242\|20 | ···················· | ···················· | ···················· | ···················· | ···················· | ···················· |
| M | 2330 hCoV-19/England/QEUH-9FB03C/2020\|EPI_ISL_588243\|20 | ···················· | ···················· | ···················· | ···················· | ···················· | ···················· |
| M | 2331 hCoV-19/England/QEUH-9FA49B/2020\|EPI_ISL_588244\|20 | ···················· | ···················· | ···················· | ···················· | ···················· | ···················· |
| M | 2332 hCoV-19/England/QEUH-9FA45F/2020\|EPI_ISL_588247\|20 | ···················· | ···················· | ···················· | ···················· | ···················· | ···················· |
| M | 2333 hCoV-19/England/QEUH-9FA589/2020\|EPI_ISL_588248\|20 | ···················· | ···················· | ···················· | ···················· | ···················· | ···················· |
| M | 2334 hCoV-19/England/QEUH-9FAEF7/2020\|EPI_ISL_588250\|20 | ···················· | ···················· | ···················· | ···················· | ···················· | ···················· |
| M | 2335 hCoV-19/England/QEUH-9FA7CF/2020\|EPI_ISL_588251\|20 | ···················· | ···················· | ···················· | ···················· | ···················· | ···················· |
| M | 2336 hCoV-19/England/QEUH-9FAF30/2020\|EPI_ISL_588253\|20 | ···················· | ···················· | ···················· | ···················· | ···················· | ···················· |
| M | 2337 hCoV-19/England/QEUH-9FABD2/2020\|EPI_ISL_588254\|20 | ···················· | ···················· | ···················· | ···················· | ···················· | ···················· |
| M | 2338 hCoV-19/England/QEUH-9FA668/2020\|EPI_ISL_588255\|20 | ···················· | ···················· | ···················· | ···················· | ···················· | ···················· |
| M | 2339 hCoV-19/England/QEUH-9FAF7C/2020\|EPI_ISL_588256\|20 | ···················· | ···················· | ···················· | ···················· | ···················· | ···················· |
| M | 2340 hCoV-19/England/QEUH-9FB588/2020\|EPI_ISL_588257\|20 | ···················· | ···················· | ···················· | ···················· | ···················· | ···················· |

| | | | | | | | | | |
|---|---|---|---|---|---|---|---|---|---|
| M | 2341 | hCoV-19/England/QEUH-9FA71A/2020\|EPI_ISL_588258\|20 | ···················· | ···················· | ···················· | ···················· | ···················· | ···················· | |
| M | 2342 | hCoV-19/England/QEUH-9FAD36/2020\|EPI_ISL_588259\|20 | ···················· | ···················· | ···················· | ···················· | ···················· | ···················· | |
| M | 2343 | hCoV-19/England/QEUH-9FA817/2020\|EPI_ISL_588260\|20 | ···················· | ···················· | ···················· | ···················· | ···················· | ···················· | |
| M | 2344 | hCoV-19/England/QEUH-9FAC2A/2020\|EPI_ISL_588261\|20 | ···················· | ···················· | ···················· | ···················· | ···················· | ···················· | |
| M | 2345 | hCoV-19/England/QEUH-9FA404/2020\|EPI_ISL_588262\|20 | ···················· | ···················· | ···················· | ···················· | ···················· | ···················· | |
| M | 2346 | hCoV-19/England/QEUH-9FA6E0/2020\|EPI_ISL_588263\|20 | ···················· | ···················· | ···················· | ···················· | ···················· | ···················· | |
| M | 2347 | hCoV-19/England/QEUH-9FB184/2020\|EPI_ISL_588265\|20 | ···················· | ···················· | ···················· | ···················· | ···················· | ···················· | |
| M | 2348 | hCoV-19/England/QEUH-9FA844/2020\|EPI_ISL_588266\|20 | ···················· | ···················· | ···················· | ···················· | ···················· | ···················· | |
| M | 2349 | hCoV-19/England/QEUH-9FB2BE/2020\|EPI_ISL_588267\|20 | ···················· | ···················· | ···················· | ···················· | ···················· | ···················· | |
| M | 2350 | hCoV-19/England/QEUH-9FAE60/2020\|EPI_ISL_588268\|20 | ···················· | ···················· | ···················· | ···················· | ···················· | ···················· | |
| M | 2351 | hCoV-19/England/QEUH-9FA3E9/2020\|EPI_ISL_588269\|20 | ···················· | ···················· | ···················· | ···················· | ···················· | ···················· | |
| M | 2352 | hCoV-19/England/QEUH-9FAE9D/2020\|EPI_ISL_588270\|20 | ···················· | ···················· | ···················· | ···················· | ···················· | ···················· | |
| M | 2353 | hCoV-19/England/QEUH-9FA826/2020\|EPI_ISL_588271\|20 | ···················· | ···················· | ···················· | ···················· | ···················· | ···················· | |
| M | 2354 | hCoV-19/England/QEUH-9FA46E/2020\|EPI_ISL_588272\|20 | ···················· | ···················· | ···················· | ···················· | ···················· | ···················· | |
| M | 2355 | hCoV-19/England/QEUH-9FAFA9/2020\|EPI_ISL_588273\|20 | ···················· | ···················· | ···················· | ···················· | ···················· | ···················· | |
| M | 2356 | hCoV-19/England/QEUH-9FB333/2020\|EPI_ISL_588274\|20 | ···················· | ···················· | ···················· | ···················· | ···················· | ···················· | |
| M | 2357 | hCoV-19/England/QEUH-9FA8DB/2020\|EPI_ISL_588275\|20 | ···················· | ···················· | ···················· | ···················· | ···················· | ···················· | |
| M | 2358 | hCoV-19/England/QEUH-9F5BE6/2020\|EPI_ISL_588276\|20 | ···················· | ···················· | ···················· | ···················· | ···················· | ···················· | |
| M | 2359 | hCoV-19/England/QEUH-9F4B14/2020\|EPI_ISL_588277\|20 | ···················· | ···················· | ···················· | ···················· | ···················· | ···················· | |
| M | 2360 | hCoV-19/England/QEUH-9F4E57/2020\|EPI_ISL_588278\|20 | ···················· | ···················· | ···················· | ···················· | ···················· | ···················· | |
| M | 2361 | hCoV-19/England/QEUH-9F5858/2020\|EPI_ISL_588279\|20 | ···················· | C··················· | ···················· | ···················· | ···················· | ···················· | |
| M | 2362 | hCoV-19/England/QEUH-9F5E92/2020\|EPI_ISL_588282\|20 | ···················· | ···················· | ···················· | ···················· | ···················· | ···················· | |
| M | 2363 | hCoV-19/England/QEUH-9F4965/2020\|EPI_ISL_588283\|20 | ···················· | ···················· | ···················· | ···················· | ····T··············· | ···················· | |
| M | 2364 | hCoV-19/Wales/QEUH-9F49FC/2020\|EPI_ISL_588284\|2020 | ···················· | ···················· | ···················· | ···················· | ···················· | ···················· | |
| M | 2365 | hCoV-19/England/QEUH-9F4B8D/2020\|EPI_ISL_588286\|20 | ···················· | ···················· | ···················· | ···················· | ···················· | ···················· | |
| M | 2366 | hCoV-19/England/QEUH-9F4947/2020\|EPI_ISL_588287\|20 | ···················· | ···················· | ···················· | ···················· | ···················· | ···················· | |
| M | 2367 | hCoV-19/England/QEUH-9F6AE8/2020\|EPI_ISL_588288\|20 | ···················· | ···················· | ···················· | ···················· | ···················· | ···················· | |
| M | 2368 | hCoV-19/England/QEUH-9F5D59/2020\|EPI_ISL_588289\|20 | ···················· | ···················· | ···················· | ···················· | ···················· | ···················· | |
| M | 2369 | hCoV-19/England/ALDP-9ECE07/2020\|EPI_ISL_589536\|20 | ···················· | ···················· | ···················· | ···················· | ···················· | ·R·················· | |
| M | 2370 | hCoV-19/England/ALDP-9ECF31/2020\|EPI_ISL_589537\|20 | ···················· | ···················· | ···················· | ···················· | ···················· | ···················· | |
| M | 2371 | hCoV-19/England/ALDP-9ED57A/2020\|EPI_ISL_589538\|20 | ···················· | ···················A | ···················· | ···················· | ···················· | ···················· | 1 |
| M | 2372 | hCoV-19/England/ALDP-9E5994/2020\|EPI_ISL_589539\|20 | ···················· | ······A············· | ···················· | ···················· | ···················· | ···················· | |
| M | 2373 | hCoV-19/England/ALDP-9E59FE/2020\|EPI_ISL_589540\|20 | ···················· | ···················· | ···················· | ···················· | ···················· | ···················· | |
| M | 2374 | hCoV-19/England/ALDP-9EDFE5/2020\|EPI_ISL_589541\|20 | ···················· | ···················· | ···················· | ···················· | ···················· | ···················· | |
| M | 2375 | hCoV-19/England/ALDP-9E5ABF/2020\|EPI_ISL_589542\|20 | ···················· | ···················· | ···················· | ···················· | ···················· | ···················· | |
| M | 2376 | hCoV-19/England/ALDP-9ED54D/2020\|EPI_ISL_589545\|20 | ···················· | ···················· | ···················· | ···················· | ···················· | ···················· | |
| M | 2377 | hCoV-19/England/ALDP-9E5AA0/2020\|EPI_ISL_589547\|20 | ···················· | ···················· | ···················· | ···················· | ···················· | ···················· | |
| M | 2378 | hCoV-19/England/ALDP-9ECE43/2020\|EPI_ISL_589548\|20 | ···················· | ···················· | ···················· | ···················· | ···················· | ···················· | |
| M | 2379 | hCoV-19/England/ALDP-9ED0F1/2020\|EPI_ISL_589549\|20 | ···················· | ···················· | ···················· | ···················· | ···················· | ···················· | |
| M | 2380 | hCoV-19/England/ALDP-9EDEBB/2020\|EPI_ISL_589550\|20 | ···················· | ···················· | ···················· | ···················· | ···················· | ···················· | |
| M | 2381 | hCoV-19/England/ALDP-9ECEAD/2020\|EPI_ISL_589552\|20 | ···················· | ···················· | ···················· | ···················· | ···················· | ···················· | |
| M | 2382 | hCoV-19/England/ALDP-9E5C9B/2020\|EPI_ISL_589553\|20 | ···················· | ···················· | ···················· | ···················· | ···················· | ···················· | |
| M | 2383 | hCoV-19/England/ALDP-9EDE9D/2020\|EPI_ISL_589554\|20 | ···················· | ···················· | ···················· | ···················· | ···················· | ···················· | |
| M | 2384 | hCoV-19/England/ALDP-9ED826/2020\|EPI_ISL_589555\|20 | ···················· | ···················· | ···················· | ···················· | ···················· | ···················· | |
| M | 2385 | hCoV-19/England/ALDP-9E5C5F/2020\|EPI_ISL_589556\|20 | ···················· | ···················· | ···················· | ···················· | ···················· | ···················· | |
| M | 2386 | hCoV-19/England/ALDP-9EE0D2/2020\|EPI_ISL_589557\|20 | ···················· | ···················· | ···················· | ···················· | ···················· | ···················· | |
| M | 2387 | hCoV-19/England/ALDP-9ED63B/2020\|EPI_ISL_589570\|20 | ···················· | ···················· | ···················· | ···················· | ···················· | ···················· | |
| M | 2388 | hCoV-19/England/ALDP-9E581F/2020\|EPI_ISL_589571\|20 | ···················· | ···················· | ···················· | ···················· | ···················· | ···················· | |
| M | 2389 | hCoV-19/England/MILK-9F0CE8/2020\|EPI_ISL_589573\|20 | ···················· | ···················· | ···················· | ···················· | ···················· | ···················· | |
| M | 2390 | hCoV-19/England/MILK-9F08A8/2020\|EPI_ISL_589574\|20 | ···················· | ···················· | ···················· | ···················· | ···················· | ···················· | |
| M | 2391 | hCoV-19/England/MILK-9F1DE4/2020\|EPI_ISL_589575\|20 | ···················· | ···················· | ···················· | ···················· | ···················· | ···················· | |
| M | 2392 | hCoV-19/England/MILK-9F0978/2020\|EPI_ISL_589576\|20 | ···················· | ···················· | ···················· | ···················· | ···················· | ···················· | |
| M | 2393 | hCoV-19/Wales/MILK-9F128B/2020\|EPI_ISL_589577\|2020 | ···················· | ···················· | ···················· | ···················· | ···················· | ···················· | |
| M | 2394 | hCoV-19/England/MILK-9F0AA2/2020\|EPI_ISL_589578\|20 | ···················· | ···················· | ···················· | ···················· | ···················· | ···················· | |
| M | 2395 | hCoV-19/England/MILK-9F0D4F/2020\|EPI_ISL_589579\|20 | ···················· | ···················· | ···················· | ···················· | ···················· | ···················· | |
| M | 2396 | hCoV-19/Wales/MILK-9F0D5E/2020\|EPI_ISL_589580\|2020 | ···················· | ···················· | ···················· | ···················· | ···················· | ···················· | |
| M | 2397 | hCoV-19/England/MILK-9F1A74/2020\|EPI_ISL_589581\|20 | ···················· | ···················· | ···················· | ···················· | ···················· | ···················· | |
| M | 2398 | hCoV-19/England/MILK-9F0DD6/2020\|EPI_ISL_589582\|20 | ···················· | ···················· | ···················· | ···················· | ···················· | ···················· | |
| M | 2399 | hCoV-19/England/MILK-9F13A6/2020\|EPI_ISL_589583\|20 | ···················· | ···················· | ···················· | ···················· | ···················· | ···················· | |
| M | 2400 | hCoV-19/Wales/MILK-9F1BDB/2020\|EPI_ISL_589584\|2020 | ···················· | ···················· | ···················· | ···················· | ···················· | ···················· | |
| M | 2401 | hCoV-19/England/MILK-9F0FC1/2020\|EPI_ISL_589585\|20 | ···················· | ·A·················· | ···················· | ···················· | ···················· | ···················· | |
| M | 2402 | hCoV-19/England/MILK-9F135B/2020\|EPI_ISL_589586\|20 | ···················· | ···················· | ···················· | ···················· | ···················· | ···················· | |
| M | 2403 | hCoV-19/England/MILK-9F1AB0/2020\|EPI_ISL_589587\|20 | ···················· | ···················· | ···················· | ···················· | ···················· | ···················· | |
| M | 2404 | hCoV-19/England/MILK-9F1018/2020\|EPI_ISL_589588\|20 | ···················· | ·A·················· | ···················· | ···················· | ···················· | ···················· | |
| M | 2405 | hCoV-19/England/MILK-9F13C4/2020\|EPI_ISL_589589\|20 | ···················· | ···················· | ···················· | ···················· | ···················· | ···················· | |
| M | 2406 | hCoV-19/England/MILK-9F1142/2020\|EPI_ISL_589591\|20 | ···················· | ···················· | ···················· | ···················· | ···················· | ···················· | |
| M | 2407 | hCoV-19/England/MILK-9F1801/2020\|EPI_ISL_589592\|20 | ···················· | ···················· | ···················· | ···················· | ···················· | ···················· | |
| M | 2408 | hCoV-19/England/MILK-9F087B/2020\|EPI_ISL_589593\|20 | ···················· | ···················· | ···················· | ···················· | ···················· | ···················· | |
| M | 2409 | hCoV-19/England/MILK-9F1EB4/2020\|EPI_ISL_589594\|20 | ···················· | ·A·················· | ···········Y········ | ···················· | ···················· | ···················· | |
| M | 2410 | hCoV-19/England/MILK-9F09B4/2020\|EPI_ISL_589596\|20 | ···················· | ···················· | ···················· | ···················· | ···················· | ···················· | |
| M | 2411 | hCoV-19/England/MILK-9F1D7B/2020\|EPI_ISL_589597\|20 | ···················· | ···················· | ···················· | ···················· | ···················· | ···················· | |
| M | 2412 | hCoV-19/England/MILK-9F1C41/2020\|EPI_ISL_589598\|20 | ···················· | ···················· | ···················· | ···················· | ···················· | ···················· | |
| M | 2413 | hCoV-19/England/MILK-9F0A57/2020\|EPI_ISL_589599\|20 | ···················· | ···················· | ···················· | ···················· | ···················· | ···················· | |
| M | 2414 | hCoV-19/England/MILK-9F1133/2020\|EPI_ISL_589600\|20 | ···················· | ···················· | ···················· | ···················· | ···················· | ···················· | |
| M | 2415 | hCoV-19/England/MILK-9F1B71/2020\|EPI_ISL_589601\|20 | ···················· | ·A·················· | ···················· | ···················· | ···················· | ···A················ | |
| M | 2416 | hCoV-19/England/MILK-9F124F/2020\|EPI_ISL_589602\|20 | ···················· | ···················· | ···················· | ···················· | ···················· | ···················· | |
| M | 2417 | hCoV-19/England/MILK-9EF4F3/2020\|EPI_ISL_589603\|20 | ···················· | ···················· | ···················· | ···················· | ···················· | ···················· | |
| M | 2418 | hCoV-19/England/MILK-9F0C9D/2020\|EPI_ISL_589604\|20 | ···················· | ···················· | ···················· | ···················· | ···················· | ···················· | |
| M | 2419 | hCoV-19/England/MILK-9F0987/2020\|EPI_ISL_589605\|20 | ···················· | ···················· | ···················· | ···················· | ···················· | ···················· | |
| M | 2420 | hCoV-19/England/MILK-9F1B80/2020\|EPI_ISL_589606\|20 | ···················· | ···················· | ···················· | ···················· | ···················· | ···················· | |

| | | | | | | | |
|---|---|---|---|---|---|---|---|
| M | 2421 hCoV-19/Wales/MILK-9F0ED3/2020\|EPI_ISL_589607\|2020 | ···················· | ···················· | ····················· | ···················· | ···················· | ···················· |
| M | 2422 hCoV-19/England/MILK-9F086C/2020\|EPI_ISL_589609\|20 | ···················· | ···················· | ····················· | ···················· | ···················· | ···················· |
| M | 2423 hCoV-19/England/MILK-9F1DC6/2020\|EPI_ISL_589610\|20 | ···················· | ···················· | ····················· | ···················· | ···················· | ···················· |
| M | 2424 hCoV-19/England/MILK-9F1AA1/2020\|EPI_ISL_589611\|20 | ···················· | ···················· | ····················· | ···················· | ···················· | ···················· |
| M | 2425 hCoV-19/England/MILK-9F0A48/2020\|EPI_ISL_589612\|20 | ···················· | ···················· | ····················· | ···················· | ···················· | ···················· |
| M | 2426 hCoV-19/England/QEUH-9B7F20/2020\|EPI_ISL_590520\|20 | ···················· | ·A·················· | ····················· | ···················· | ···················· | ···················· |
| M | 2427 hCoV-19/England/QEUH-9B7315/2020\|EPI_ISL_590521\|20 | ···················· | ···················· | ····················· | ···················· | ···················· | ···················· |
| M | 2428 hCoV-19/England/QEUH-9B7096/2020\|EPI_ISL_590522\|20 | ···················· | ···················· | ····················· | ···················· | ···················· | ···················· |
| M | 2429 hCoV-19/England/QEUH-9B79AA/2020\|EPI_ISL_590524\|20 | ···················· | ···················· | ····················· | ···················· | ···················· | ···················· |
| M | 2430 hCoV-19/England/QEUH-9B752E/2020\|EPI_ISL_590525\|20 | ···················· | ···················· | ····················· | ···················· | ···················· | ···················· |
| M | 2431 hCoV-19/England/QEUH-9B8138/2020\|EPI_ISL_590527\|20 | ···················· | ···················· | ····················· | ···················· | ······T············· | ···················· |
| M | 2432 hCoV-19/England/QEUH-9B744F/2020\|EPI_ISL_590528\|20 | ···················· | ···················· | ····················· | ···················· | ···················· | ···················· |
| M | 2433 hCoV-19/England/QEUH-9B7166/2020\|EPI_ISL_590529\|20 | ···················· | ···················· | ····················· | ···················· | ···················· | ···················· |
| M | 2434 hCoV-19/England/QEUH-9B7B86/2020\|EPI_ISL_590530\|20 | ···················· | ·A·················· | ····················· | ···················· | ···················· | ···A················ |
| M | 2435 hCoV-19/England/QEUH-9B738E/2020\|EPI_ISL_590531\|20 | ···················· | ···················· | ····················· | ···················· | ···················· | ···················· |
| M | 2436 hCoV-19/England/QEUH-9B77BF/2020\|EPI_ISL_590532\|20 | ···················· | ···················· | ····················· | ···················· | ···················· | ···················· |
| M | 2437 hCoV-19/England/QEUH-9B8156/2020\|EPI_ISL_590533\|20 | ···················· | ···················· | ····················· | ···················· | ···················· | ···················· |
| M | 2438 hCoV-19/England/QEUH-9B7FC6/2020\|EPI_ISL_590534\|20 | ···················· | ···················· | ····················· | ···················· | ···················· | ···················· |
| M | 2439 hCoV-19/England/QEUH-9B7FD5/2020\|EPI_ISL_590535\|20 | ···················· | ···················· | ····················· | ···················· | ···················· | ···················· |
| M | 2440 hCoV-19/England/QEUH-9B746D/2020\|EPI_ISL_590536\|20 | ···················· | ·A·················· | ····················· | ···················· | ···················· | ···················· |
| M | 2441 hCoV-19/England/QEUH-9B700F/2020\|EPI_ISL_590537\|20 | ···················· | ···················· | ····················· | ···················· | ···················· | ···················· |
| M | 2442 hCoV-19/England/QEUH-9B7F7B/2020\|EPI_ISL_590538\|20 | ···················· | ···················· | ····················· | ···················· | ····T··············· | ···················· |
| M | 2443 hCoV-19/England/QEUH-9B8165/2020\|EPI_ISL_590539\|20 | ···················· | ···················· | ····················· | ···················· | ···················· | ···················· |
| M | 2444 hCoV-19/England/QEUH-9B7139/2020\|EPI_ISL_590545\|20 | ···················· | ·A·················· | ····················· | ···················· | ···················· | ···A················ |
| M | 2445 hCoV-19/England/QEUH-9B8068/2020\|EPI_ISL_590547\|20 | ···················· | ···················· | ····················· | ···················· | ···················· | ···················· |
| M | 2446 hCoV-19/England/QEUH-9B7AF2/2020\|EPI_ISL_590548\|20 | ···················· | ···················· | ····················· | ···················· | ···················· | ···················· |
| M | 2447 hCoV-19/England/QEUH-9B7333/2020\|EPI_ISL_590550\|20 | ···················· | ·A·················· | ····················· | ···················· | ···················· | ···A················ |
| M | 2448 hCoV-19/England/QEUH-9B78F8/2020\|EPI_ISL_590551\|20 | ···················· | ···················· | ····················· | ···················· | ······T············· | ···················· |
| M | 2449 hCoV-19/England/QEUH-9B762B/2020\|EPI_ISL_590552\|20 | ···················· | ···················· | ····················· | ···················· | ···················· | ···················· |
| M | 2450 hCoV-19/England/QEUH-9B6F4F/2020\|EPI_ISL_590553\|20 | ···················· | ···················· | ····················· | ···················· | ···················· | ···················· |
| M | 2451 hCoV-19/England/QEUH-9B7825/2020\|EPI_ISL_590554\|20 | ···················· | ·A·················· | ····················· | ···················· | ···················· | ···················· |
| M | 2452 hCoV-19/England/QEUH-9B7F02/2020\|EPI_ISL_590555\|20 | ···················· | ···················· | ····················· | ···················· | ···················· | ···················· |
| M | 2453 hCoV-19/England/QEUH-9B7DAE/2020\|EPI_ISL_590556\|20 | ···················· | ···················· | ····················· | ···················· | ···················· | ···················· |
| M | 2454 hCoV-19/England/QEUH-9B7FA8/2020\|EPI_ISL_590557\|20 | ···················· | ···················· | ····················· | ···················· | ···················· | ···················· |
| M | 2455 hCoV-19/England/QEUH-9B755B/2020\|EPI_ISL_590558\|20 | ···················· | ···················· | ····················· | ···················· | ···················· | ···················· |
| M | 2456 hCoV-19/England/QEUH-9B7EAB/2020\|EPI_ISL_590559\|20 | ···················· | ···················· | ····················· | ···················· | ···················· | ···················· |
| M | 2457 hCoV-19/England/QEUH-9B7D53/2020\|EPI_ISL_590560\|20 | ···················· | ···················· | ····················· | ···················· | ···················· | ···················· |
| M | 2458 hCoV-19/England/QEUH-9B72CD/2020\|EPI_ISL_590562\|20 | ···················· | ···················· | ····················· | ···················· | ···················· | ···················· |
| M | 2459 hCoV-19/England/QEUH-9B7E6F/2020\|EPI_ISL_590565\|20 | ···················· | ···················· | ····················· | ···················· | ···················· | ···················· |
| M | 2460 hCoV-19/England/QEUH-9B79C8/2020\|EPI_ISL_590566\|20 | ···················· | ···················· | ····················· | ···················· | ···················· | ···················· |
| M | 2461 hCoV-19/England/QEUH-9B79D7/2020\|EPI_ISL_590567\|20 | ···················· | ···················· | ····················· | ···················· | ···················· | ···················· |
| M | 2462 hCoV-19/England/QEUH-9B7193/2020\|EPI_ISL_590568\|20 | ···················· | ···················· | ····················· | ···················· | ···················· | ···················· |
| M | 2463 hCoV-19/England/QEUH-9B7719/2020\|EPI_ISL_590569\|20 | ···················· | ···················· | ····················· | ···················· | ···················· | ···················· |
| M | 2464 hCoV-19/England/QEUH-9B7A4D/2020\|EPI_ISL_590571\|20 | ···················· | ···················· | ····················· | ···················· | ···················· | ···················· |
| M | 2465 hCoV-19/England/QEUH-9B7DEA/2020\|EPI_ISL_590572\|20 | ···················· | ···················· | ····················· | ···················· | ···················· | ···················· |
| M | 2466 hCoV-19/England/QEUH-9B6FF4/2020\|EPI_ISL_590573\|20 | ···················· | ···················· | ····················· | ···················· | ···················· | ···················· |
| M | 2467 hCoV-19/England/QEUH-9B6F6D/2020\|EPI_ISL_590574\|20 | ···················· | ···················· | ····················· | ···················· | ···················· | ···················· |
| M | 2468 hCoV-19/England/QEUH-9B75B5/2020\|EPI_ISL_590575\|20 | ···················· | ·A·················· | ····················· | ···················· | ···················· | ···················· |
| M | 2469 hCoV-19/England/QEUH-9B704B/2020\|EPI_ISL_590577\|20 | ···················· | ···················· | ····················· | ···················· | ···················· | ···················· |
| M | 2470 hCoV-19/England/QEUH-9B7D26/2020\|EPI_ISL_590578\|20 | ···················· | ···················· | ····················· | ···················· | ···················· | ···················· |
| M | 2471 hCoV-19/England/QEUH-9B7782/2020\|EPI_ISL_590579\|20 | ···················· | ·A·················· | ····················· | ···················· | ···················· | ···················· |
| M | 2472 hCoV-19/England/QEUH-9B7C38/2020\|EPI_ISL_590580\|20 | ···················· | ···················· | ····················· | ···················· | ···················· | ···················· |
| M | 2473 hCoV-19/England/QEUH-9B7FF3/2020\|EPI_ISL_590581\|20 | ···················· | ···················· | ····················· | ···················· | ···················· | ···················· |
| M | 2474 hCoV-19/England/QEUH-9B7236/2020\|EPI_ISL_590582\|20 | ···················· | ···················· | ····················· | ···················· | ···················· | ···················· |
| M | 2475 hCoV-19/England/QEUH-9B6ED9/2020\|EPI_ISL_590583\|20 | ···················· | ···················· | ····················· | ···················· | ···················· | ···················· |
| M | 2476 hCoV-19/England/QEUH-9B751F/2020\|EPI_ISL_590584\|20 | ···················· | ···················· | ····················· | ···················· | ···················· | ···················· |
| M | 2477 hCoV-19/Wales/QEUH-9B756A/2020\|EPI_ISL_590585\|2020 | ···················· | ···················· | ····················· | ···················· | ···················· | ···················· |
| M | 2478 hCoV-19/England/QEUH-9B7A89/2020\|EPI_ISL_590586\|20 | ···················· | ···················· | ····················· | ···················· | ···················· | ···················· |
| M | 2479 hCoV-19/England/QEUH-9B7E32/2020\|EPI_ISL_590587\|20 | ···················· | ···················· | ····················· | ···················· | ···················· | ···················· |
| M | 2480 hCoV-19/England/QEUH-9B7816/2020\|EPI_ISL_590588\|20 | ···················· | ···················· | ····················· | ···················· | ····T··············· | ···················· |
| M | 2481 hCoV-19/England/QEUH-9B7272/2020\|EPI_ISL_590589\|20 | ···················· | ···················· | ····················· | ···················· | ···················· | ···················· |
| M | 2482 hCoV-19/England/QEUH-9B7BB3/2020\|EPI_ISL_590590\|20 | ···················· | ···················· | ····················· | ···················· | ···················· | ···················· |
| M | 2483 hCoV-19/England/QEUH-9B7870/2020\|EPI_ISL_590591\|20 | ···················· | ···················· | ····················· | ···················· | ···················· | ···················· |
| M | 2484 hCoV-19/England/QEUH-9B811A/2020\|EPI_ISL_590592\|20 | ···················· | ···················· | ····················· | ···················· | ···················· | ···················· |
| M | 2485 hCoV-19/NorthernIreland/QEUH-9B70C3/2020\|EPI_ISL_5 | ···················· | ···················· | ····················· | ···················· | ···················· | ···················· |
| M | 2486 hCoV-19/England/QEUH-9B7CED/2020\|EPI_ISL_590596\|20 | ···················· | ···················· | ····················· | ···················· | ····T··············· | ···················· |
| M | 2487 hCoV-19/England/QEUH-9B80B3/2020\|EPI_ISL_590597\|20 | ···················· | ···················· | ····················· | ···················· | ···················· | ···················· |
| M | 2488 hCoV-19/England/QEUH-9B7AB6/2020\|EPI_ISL_590598\|20 | ···················· | ···················· | ····················· | ···················· | ···················· | ···················· |
| M | 2489 hCoV-19/England/QEUH-9B737F/2020\|EPI_ISL_590599\|20 | ···················· | ···················· | ····················· | ···················· | ···················· | ···················· |
| M | 2490 hCoV-19/Australia/VIC11782/2020\|EPI_ISL_591850\|202 | ···················· | ···················· | ····················· | ···················· | ···················· | ···················· |
| M | 2491 hCoV-19/Australia/VIC11784/2020\|EPI_ISL_591851\|202 | ···················· | ···················· | ····················· | ···················· | ···················· | ···················· |
| M | 2492 hCoV-19/Australia/VIC11787/2020\|EPI_ISL_591852\|202 | ···················· | ···················· | ····················· | ···················· | ···················· | ···················· |
| M | 2493 hCoV-19/Australia/VIC11791/2020\|EPI_ISL_591853\|202 | ···················· | ···················· | ····················· | ···················· | ···················· | ···················· |
| M | 2494 hCoV-19/Australia/VIC11797/2020\|EPI_ISL_591854\|202 | ···················· | ···················· | ····················· | ···················· | ···················· | ···················· |
| M | 2495 hCoV-19/Australia/VIC11798/2020\|EPI_ISL_591855\|202 | ···················· | ···················· | ···K················· | ···················· | ···················· | ···················· |
| M | 2496 hCoV-19/Australia/VIC11800/2020\|EPI_ISL_591856\|202 | ···················· | ···················· | ····················· | ···················· | ···················· | ···················· |
| M | 2497 hCoV-19/Australia/VIC11801/2020\|EPI_ISL_591857\|202 | ···················· | ···················· | ····················· | ···················· | ···················· | ···················· |
| M | 2498 hCoV-19/Australia/VIC11803/2020\|EPI_ISL_591858\|202 | ···················· | ···················· | ····················· | ···················· | ···················· | ···················· |
| M | 2499 hCoV-19/Australia/VIC11810/2020\|EPI_ISL_591859\|202 | ···················· | ···················· | ···K················· | ···················· | ···················· | ···················· |
| M | 2500 hCoV-19/Australia/VIC11811/2020\|EPI_ISL_591860\|202 | ···················· | ···················· | ····················· | ···················· | ···················· | ···················· |

| | | | | | | | | |
|---|---|---|---|---|---|---|---|---|
| M | 2501 | hCoV-19/Australia/VIC11817/2020\|EPI_ISL_591861\|202 | ···················· | ···················· | ····················· | ···················· | ···················· | ···················· |
| M | 2502 | hCoV-19/Australia/VIC11819/2020\|EPI_ISL_591862\|202 | ···················· | ···················· | ····················· | ···················· | ···················· | ···················· |
| M | 2503 | hCoV-19/Australia/VIC11820/2020\|EPI_ISL_591863\|202 | ···················· | ···················· | ····················· | ···················· | ···················· | ···················· |
| M | 2504 | hCoV-19/Australia/VIC11822/2020\|EPI_ISL_591864\|202 | ···················· | ···················· | ···C················· | ···················· | ···················· | ···················· |
| M | 2505 | hCoV-19/Australia/VIC11824/2020\|EPI_ISL_591865\|202 | ···················· | ···················· | ···K················· | ···················· | ···················· | ···················· |
| M | 2506 | hCoV-19/Australia/VIC11826/2020\|EPI_ISL_591866\|202 | ···················· | ···················· | ····················· | ···················· | ···················· | ···················· |
| M | 2507 | hCoV-19/Australia/VIC11827/2020\|EPI_ISL_591867\|202 | ···················· | ···················· | ····················· | ···················· | ···················· | ···················· |
| M | 2508 | hCoV-19/Australia/VIC11829/2020\|EPI_ISL_591868\|202 | ···················· | ···················· | ···K················· | ···················· | ···················· | ···················· |
| M | 2509 | hCoV-19/Australia/VIC11830/2020\|EPI_ISL_591869\|202 | ···················· | ···················· | ···K················· | ···················· | ···················· | ···················· |
| M | 2510 | hCoV-19/Australia/VIC11867/2020\|EPI_ISL_591876\|202 | ···················· | ···················· | ····················· | ···················· | ···················· | ···················· |
| M | 2511 | hCoV-19/Australia/VIC11871/2020\|EPI_ISL_591878\|202 | ···················· | ···················· | ····················· | ···················· | ···················· | ···················· |
| M | 2512 | hCoV-19/Australia/VIC11872/2020\|EPI_ISL_591879\|202 | ···················· | ···················· | ····················· | ···················· | ···················· | ···················· |
| M | 2513 | hCoV-19/Australia/VIC11877/2020\|EPI_ISL_591880\|202 | ···················· | ···················· | ···K················· | ···················· | ···················· | ···················· |
| M | 2514 | hCoV-19/Australia/VIC11878/2020\|EPI_ISL_591881\|202 | ···················· | ···················· | ····················· | ···················· | ···················· | ···················· |
| M | 2515 | hCoV-19/Australia/VIC11879/2020\|EPI_ISL_591882\|202 | ···················· | ···················· | ···K················· | ···················· | ···················· | ···················· |
| M | 2516 | hCoV-19/Australia/VIC11881/2020\|EPI_ISL_591883\|202 | ···················· | ···················· | ····················· | ···················· | ···················· | ···················· |
| M | 2517 | hCoV-19/Australia/VIC11890/2020\|EPI_ISL_591884\|202 | ···················· | ···················· | ····················· | ···················· | ···················· | ···················· |
| M | 2518 | hCoV-19/Australia/VIC11891/2020\|EPI_ISL_591885\|202 | ···················· | ···················· | ···K················· | ···················· | ···················· | ···················· |
| M | 2519 | hCoV-19/Australia/VIC11897/2020\|EPI_ISL_591886\|202 | ···········N········ | ···················· | ···K················· | ···················· | ···················· | ···················· |
| M | 2520 | hCoV-19/Australia/VIC11899/2020\|EPI_ISL_591887\|202 | ···················· | ···················· | ····················· | ···················· | ···················· | ···················· |
| M | 2521 | hCoV-19/Australia/VIC11901/2020\|EPI_ISL_591888\|202 | ···················· | ···················· | ···K················· | ···················· | ···················· | ···················· |
| M | 2522 | hCoV-19/Australia/VIC11902/2020\|EPI_ISL_591889\|202 | ···················· | ···················· | ····················· | ···················· | ···················· | ···················· |
| M | 2523 | hCoV-19/Australia/VIC11906/2020\|EPI_ISL_591890\|202 | ···················· | ···················· | ····················· | ···················· | ···················· | ···················· |
| M | 2524 | hCoV-19/Australia/VIC11909/2020\|EPI_ISL_591891\|202 | ···················· | ···················· | ····················· | ···················· | ···················· | ···················· |
| M | 2525 | hCoV-19/Australia/VIC11912/2020\|EPI_ISL_591892\|202 | ···················· | ···················· | ···K················· | ···················· | ···················· | ···················· |
| M | 2526 | hCoV-19/Australia/VIC11913/2020\|EPI_ISL_591893\|202 | ···················· | ···················· | ···K················· | ···················· | ···················· | ···················· |
| M | 2527 | hCoV-19/Australia/VIC11914/2020\|EPI_ISL_591894\|202 | ···················· | ···················· | ····················· | ···················· | ···················· | ···················· |
| M | 2528 | hCoV-19/Australia/VIC11918/2020\|EPI_ISL_591895\|202 | ···················· | ···················· | ····················· | ···················· | ···················· | ···················· |
| M | 2529 | hCoV-19/Australia/VIC11921/2020\|EPI_ISL_591896\|202 | ···················· | ···················· | ····················· | ···················· | ···················· | ···················· |
| M | 2530 | hCoV-19/Australia/VIC11934/2020\|EPI_ISL_591897\|202 | ···················· | ···················· | ····················· | ···················· | ···················· | ···················· |
| M | 2531 | hCoV-19/Australia/VIC11935/2020\|EPI_ISL_591898\|202 | ···················· | ···················· | ···K················· | ···················· | ···················· | ···················· |
| M | 2532 | hCoV-19/Australia/VIC11942/2020\|EPI_ISL_591899\|202 | ···················· | ···················· | ····················· | ···················· | ······R············· | ···················· |
| M | 2533 | hCoV-19/Australia/VIC11988/2020\|EPI_ISL_591906\|202 | ···················· | ···················· | ····················· | ···················· | ···················· | ···················· |
| M | 2534 | hCoV-19/Australia/VIC11994/2020\|EPI_ISL_591907\|202 | ···················· | ···················· | ····················· | ···················· | ···················· | ···················· |
| M | 2535 | hCoV-19/Australia/VIC12056/2020\|EPI_ISL_591910\|202 | ···················· | ···················· | ····················· | ···················· | ···················· | ···················· |
| M | 2536 | hCoV-19/Australia/VIC12059/2020\|EPI_ISL_591911\|202 | ···················· | ···················· | ····················· | ···················· | ···················· | ···················· |
| M | 2537 | hCoV-19/Australia/VIC12082/2020\|EPI_ISL_591912\|202 | ···················· | ···················· | ····················· | ···················· | ···················· | ···················· |
| M | 2538 | hCoV-19/Australia/VIC12083/2020\|EPI_ISL_591913\|202 | ···················· | ···················· | ····················· | ···················· | ···················· | ···················· |
| M | 2539 | hCoV-19/Australia/VIC12085/2020\|EPI_ISL_591914\|202 | ···················· | ···················· | ····················· | ···················· | ···················· | ···················· |
| M | 2540 | hCoV-19/Australia/VIC12089/2020\|EPI_ISL_591915\|202 | ···················· | ···················· | ····················· | ···················· | ···················· | ···················· |
| M | 2541 | hCoV-19/Australia/VIC12107/2020\|EPI_ISL_591916\|202 | ···················· | ···················· | ····················· | ···················· | ···················· | ···················· |
| M | 2543 | hCoV-19/Australia/VIC12120/2020\|EPI_ISL_591918\|202 | ···················· | ···················· | ····················· | ···················· | ···················· | ···················· |
| M | 2544 | hCoV-19/Australia/VIC12125/2020\|EPI_ISL_591919\|202 | ···················· | ···················· | ····················· | ···················· | ···················· | ···················· |
| M | 2546 | hCoV-19/Australia/VIC12148/2020\|EPI_ISL_591921\|202 | ···················· | ···················· | ····················· | ···················· | ···················· | ···················· |
| M | 2547 | hCoV-19/Australia/VIC12175/2020\|EPI_ISL_591922\|202 | ···················· | ···················· | ····················· | ···················· | ···················· | ···················· |
| M | 2548 | hCoV-19/Australia/VIC12182/2020\|EPI_ISL_591923\|202 | ···················· | ···················· | ····················· | ···················· | ···················· | ···················· |
| M | 2549 | hCoV-19/Australia/VIC12189/2020\|EPI_ISL_591924\|202 | ···················· | ···················· | ····················· | ···················· | ···················· | ···················· |
| M | 2550 | hCoV-19/Australia/VIC12193/2020\|EPI_ISL_591925\|202 | ···················· | ···················· | ····················· | ···················· | ···················· | ···················· |
| M | 2551 | hCoV-19/Australia/VIC12199/2020\|EPI_ISL_591926\|202 | ···················· | ···················· | ····················· | ···················· | ···················· | ···················· |
| M | 2552 | hCoV-19/Australia/VIC12202/2020\|EPI_ISL_591927\|202 | ···················· | ···················· | ····················· | ···················· | ···················· | ···················· |
| M | 2553 | hCoV-19/Australia/VIC12203/2020\|EPI_ISL_591928\|202 | ···················· | ···················· | ····················· | ···················· | ···················· | ···················· |
| M | 2554 | hCoV-19/Australia/VIC12204/2020\|EPI_ISL_591929\|202 | ···················· | ···················· | ····················· | ···················· | ···················· | ···················· |
| M | 2555 | hCoV-19/Australia/WA251/2020\|EPI_ISL_594167\|2020-0 | ···················· | ···················· | ····················· | ···················· | ···················· | ···················· |
| M | 2556 | hCoV-19/Australia/WA256/2020\|EPI_ISL_594170\|2020-0 | ···················· | ···················· | ····················· | ···················· | ···················· | ···················· |
| M | 2557 | hCoV-19/Australia/WA257/2020\|EPI_ISL_594171\|2020-0 | ···················· | ···················· | ····················· | ···················· | ···················· | ···················· |
| M | 2558 | hCoV-19/Australia/WA261/2020\|EPI_ISL_594174\|2020-0 | ···················· | ···················· | ····················· | ···················· | ···················· | ···················· |
| M | 2559 | hCoV-19/Australia/WA263/2020\|EPI_ISL_594175\|2020-0 | ···················· | ···················· | ····················· | ···················· | ···················· | ···················· |
| M | 2560 | hCoV-19/Wales/PHWC-47E77D/2020\|EPI_ISL_595498\|2020 | ···················· | ···················· | ····················· | ···················· | ···················· | ···················· |
| M | 2561 | hCoV-19/Brazil/SP-359/2020\|EPI_ISL_583497\|2020-06- | ···················· | ···················· | ····················· | ···················· | ···················· | ···················· |
| M | 2562 | hCoV-19/Brazil/SP-360/2020\|EPI_ISL_583498\|2020-06- | ···················· | ···················· | ····················· | ···················· | ···················· | ···················· |
| M | 2563 | hCoV-19/Brazil/SP-368/2020\|EPI_ISL_583499\|2020-06- | ···················· | ···················· | ····················· | ···················· | ···················· | ···················· |
| M | 2564 | hCoV-19/England/NOTT-114B52/2020\|EPI_ISL_584763\|20 | ···················· | ···················· | ····················· | ···················· | ···················· | ···················· |
| M | 2565 | hCoV-19/England/NOTT-114B61/2020\|EPI_ISL_584764\|20 | ···················· | ···················· | ····················· | ···················· | ···················· | ···················· |
| M | 2566 | hCoV-19/England/NOTT-114C22/2020\|EPI_ISL_584765\|20 | ···················· | ···················· | ····················· | ···················· | ···················· | ···················· |
| M | 2567 | hCoV-19/England/NOTT-114C6E/2020\|EPI_ISL_584766\|20 | ···················· | ···················· | ····················· | ···················· | ···················· | ···················· |
| M | 2568 | hCoV-19/England/NOTT-114CF5/2020\|EPI_ISL_584767\|20 | ···················· | ···················· | ····················· | ···················· | ···················· | ···················· |
| M | 2569 | hCoV-19/England/NOTT-114D4D/2020\|EPI_ISL_584768\|20 | ···················· | ···················· | ····················· | ···················· | ···················· | ···················· |
| M | 2570 | hCoV-19/England/NOTT-114D6B/2020\|EPI_ISL_584769\|20 | ···················· | ···················· | ····················· | ···················· | ···················· | ···················· |
| M | 2571 | hCoV-19/England/NOTT-114D89/2020\|EPI_ISL_584770\|20 | ···················· | ···················· | ····················· | ···················· | ···················· | ···················· |
| M | 2572 | hCoV-19/England/NOTT-114DA7/2020\|EPI_ISL_584771\|20 | ···················· | ···················· | ····················· | ···················· | ···················· | ···················· |
| M | 2573 | hCoV-19/England/NOTT-114DB6/2020\|EPI_ISL_584772\|20 | ···················· | ···················· | ····················· | ···················· | ···················· | ···················· |
| M | 2574 | hCoV-19/England/NOTT-114DD4/2020\|EPI_ISL_584773\|20 | ···················· | ···················· | ····················· | ···················· | ···················· | ···················· |
| M | 2575 | hCoV-19/England/NOTT-114E0E/2020\|EPI_ISL_584774\|20 | ···················· | ···················· | ····················· | ···················· | ···················· | ···················· |
| M | 2576 | hCoV-19/England/NOTT-114ED1/2020\|EPI_ISL_584777\|20 | ···················· | ···················· | ····················· | ···················· | ···················· | ···················· |
| M | 2577 | hCoV-19/England/NOTT-114EFF/2020\|EPI_ISL_584779\|20 | ···················· | ···················· | ····················· | ···················· | ···················· | ···················· |
| M | 2578 | hCoV-19/England/NOTT-114F47/2020\|EPI_ISL_584780\|20 | ···················· | ···················· | ····················· | ···················· | ···················· | ···················· |
| M | 2579 | hCoV-19/England/NOTT-114F56/2020\|EPI_ISL_584781\|20 | ···················· | ···················· | ····················· | ···················· | ···················· | ···················· |
| M | 2580 | hCoV-19/England/NOTT-114F65/2020\|EPI_ISL_584782\|20 | ···················· | ···················· | ····················· | ···················· | ···················· | ···················· |
| M | 2581 | hCoV-19/England/NOTT-114F92/2020\|EPI_ISL_584783\|20 | ···················· | ···················· | ····················· | ···················· | ···················· | ···················· |
| M | 2582 | hCoV-19/England/NOTT-114FA1/2020\|EPI_ISL_584784\|20 | ···················· | ···················· | ····················· | ···················· | ···················· | ···················· |

```
M 2583 hCoV-19/England/NOTT-114FCF/2020|EPI_ISL_584785|20 ···················· ···················· ····················· ···················· ···················· ····················
M 2584 hCoV-19/England/NOTT-114FDE/2020|EPI_ISL_584786|20 ···················· ···················· ····················· ···················· ···················· ····················
M 2585 hCoV-19/England/NOTT-114FED/2020|EPI_ISL_584787|20 ···················· ···················· ····················· ···················· ···················· ····················
M 2586 hCoV-19/England/NOTT-1150AD/2020|EPI_ISL_584788|20 ···················· ···················· ····················· ···················· ···················· ····················
M 2587 hCoV-19/England/NOTT-115104/2020|EPI_ISL_584790|20 ···················· ···················· ····················· ···················· ···················· ····················
M 2588 hCoV-19/England/NOTT-115113/2020|EPI_ISL_584791|20 ···················· ···················· ····················· ···················· ···················· ····················
M 2589 hCoV-19/England/NOTT-115131/2020|EPI_ISL_584792|20 ···················· ···················· ····················· ···················· ···················· ····················
M 2590 hCoV-19/England/NOTT-11518C/2020|EPI_ISL_584793|20 ···················· ·A·················· ····················· ···················· ···················· ···A················
M 2591 hCoV-19/England/NOTT-11519B/2020|EPI_ISL_584794|20 ···················· ·A·················· ····················· ···················· ···················· ···A················
M 2592 hCoV-19/England/NOTT-1151AA/2020|EPI_ISL_584795|20 ···················· ···················· ····················· ···················· ···················· ····················
M 2593 hCoV-19/England/NOTT-1151B9/2020|EPI_ISL_584796|20 ···················· ···················· ····················· ···················· ···················· ····················
M 2594 hCoV-19/England/NOTT-1151C8/2020|EPI_ISL_584797|20 ···················· ···················· ····················· ···················· ···················· ····················
M 2595 hCoV-19/England/NOTT-1151E6/2020|EPI_ISL_584799|20 ···················· ···················· ····················· ···················· ···················· ····················
M 2596 hCoV-19/England/QEUH-9F47D4/2020|EPI_ISL_588301|20 ···················· ···················· ····················· ···················· ···················· ····················
M 2597 hCoV-19/England/QEUH-9F4EEE/2020|EPI_ISL_588302|20 ···················· ···················· ····················· ···················· ···················· ····················
M 2598 hCoV-19/England/QEUH-9F6A42/2020|EPI_ISL_588303|20 ···················· ···················· ····················· ···················· ···················· ····················
M 2599 hCoV-19/England/QEUH-9F4A9F/2020|EPI_ISL_588304|20 ···················· ···················· ····················· ···················· ····T··············· ····················
M 2600 hCoV-19/England/QEUH-9F5E65/2020|EPI_ISL_588306|20 ···················· ···················· ····················· ···················· ···················· ····················
M 2601 hCoV-19/England/QEUH-9F48A4/2020|EPI_ISL_588307|20 ···················· ···················· ····················· ···················· ···················· ····················
M 2602 hCoV-19/England/QEUH-9F4EA2/2020|EPI_ISL_588308|20 ···················· ···················· ····················· ···················· ···················· ····················
M 2603 hCoV-19/England/QEUH-9F57F1/2020|EPI_ISL_588309|20 ···················· ···················· ····················· ···················· ···················· ····················
M 2604 hCoV-19/England/QEUH-9F6A60/2020|EPI_ISL_588310|20 ···················· ···················· ····················· ···················· ···················· ····················
M 2605 hCoV-19/England/QEUH-9F68C0/2020|EPI_ISL_588311|20 ···················· ···················· ····················· ···················· ···················· ····················
M 2606 hCoV-19/England/QEUH-9F4E48/2020|EPI_ISL_588312|20 ···················· ···················· ····················· ···················· ···················· ····················
M 2607 hCoV-19/Northern Ireland/QEUH-9F4859/2020|EPI_ISL_ ···················· ···················· ····················· ···················· ···················· ····················
M 2608 hCoV-19/England/QEUH-9F5894/2020|EPI_ISL_588314|20 ···················· ···················· ····················· ···················· ···················· ····················
M 2609 hCoV-19/England/QEUH-9F65BA/2020|EPI_ISL_588315|20 ···················· ···················· ····················· ···················· ···················· ····················
M 2610 hCoV-19/England/QEUH-9F4C20/2020|EPI_ISL_588316|20 ···················· ···················· ····················· ···················· ···················· ····················
M 2611 hCoV-19/England/QEUH-9F4C4E/2020|EPI_ISL_588317|20 ···················· ···················· ····················· ···················· ···················· ····················
M 2612 hCoV-19/England/QEUH-9F68EE/2020|EPI_ISL_588318|20 ···················· ···················· ····················· ···················· ···················· ····················
M 2613 hCoV-19/England/QEUH-9F6AAC/2020|EPI_ISL_588319|20 ···················· ···················· ····················· ···················· ···················· ····················
M 2614 hCoV-19/England/QEUH-9F5FF9/2020|EPI_ISL_588320|20 ···················· ···················· ····················· ···················· ···················· ····················
M 2615 hCoV-19/England/QEUH-9F4C3F/2020|EPI_ISL_588322|20 ···················· ···················· ····················· ···················· ···················· ····················
M 2616 hCoV-19/England/QEUH-9F5919/2020|EPI_ISL_588323|20 ···················· ···················· ····················· ···················· ···················· ····················
M 2617 hCoV-19/England/QEUH-9F4929/2020|EPI_ISL_588324|20 ···················· ···················· ····················· ···················· ···················· ····················
M 2618 hCoV-19/England/QEUH-9F5B04/2020|EPI_ISL_588325|20 ···················· ···················· ····················· ···················· ···················· ····················
M 2619 hCoV-19/England/MILK-9F1643/2020|EPI_ISL_589613|20 ···················· ···················· ····················· ···················· ···················· ····················
M 2620 hCoV-19/England/MILK-9F192C/2020|EPI_ISL_589614|20 ···················· ···················· ····················· ···················· ···················· ····················
M 2621 hCoV-19/England/MILK-9F0A75/2020|EPI_ISL_589615|20 ···················· ···················· ····················· ···················· ···················· ····················
M 2622 hCoV-19/England/MILK-9F1C9C/2020|EPI_ISL_589616|20 ···················· ···················· ····················· ···················· ···················· ····················
M 2623 hCoV-19/England/MILK-9F1995/2020|EPI_ISL_589618|20 ···················· ···················· ····················· ···················· ···················· ····················
M 2624 hCoV-19/England/MILK-9F09F0/2020|EPI_ISL_589620|20 ···················· ···················· ····················· ···················· ···················· ····················
M 2625 hCoV-19/England/MILK-9F1986/2020|EPI_ISL_589621|20 ···················· ···················· ····················· ···················· ···················· ····················
M 2626 hCoV-19/England/MILK-9F09C3/2020|EPI_ISL_589623|20 ···················· ···················· ····················· ···················· ···················· ····················
M 2627 hCoV-19/England/MILK-9F0A1B/2020|EPI_ISL_589624|20 ···················· ···················· ····················· ···················· ···················· ····················
M 2628 hCoV-19/England/MILK-9F129A/2020|EPI_ISL_589625|20 ···················· ···················· ····················· ···················· ···················· ····················
M 2629 hCoV-19/England/MILK-9F1072/2020|EPI_ISL_589626|20 ···················· ···················· ····················· ···················· ···················· ····················
M 2630 hCoV-19/England/MILK-9F19A4/2020|EPI_ISL_589627|20 ···················· ···················· ····················· ···················· ···················· ····················
M 2631 hCoV-19/England/MILK-9F194A/2020|EPI_ISL_589628|20 ···················· ···················· ····················· ···················· ···················· ····················
M 2632 hCoV-19/England/MILK-9F193B/2020|EPI_ISL_589629|20 ···················· ·A·················· ····················· ···················· ···················· ···A················
M 2633 hCoV-19/England/MILK-9F1D4E/2020|EPI_ISL_589630|20 ···················· ···················· ····················· ···················· ···················· ····················
M 2634 hCoV-19/England/MILK-9F1494/2020|EPI_ISL_589631|20 ···················· ···················· ····················· ···················· ···················· ····················
M 2635 hCoV-19/England/MILK-9F1EE1/2020|EPI_ISL_589632|20 ···················· ···················· ····················· ···················· ······T············· ····················
M 2636 hCoV-19/England/MILK-9F1D11/2020|EPI_ISL_589633|20 ···················· ···················· ····················· ···················· ···················· ····················
M 2637 hCoV-19/England/MILK-9F084E/2020|EPI_ISL_589634|20 ···················· ···················· ····················· ···················· ···················· ····················
M 2638 hCoV-19/England/MILK-9F0969/2020|EPI_ISL_589635|20 ···················· ···················· ····················· ···················· ···················· ····················
M 2639 hCoV-19/England/MILK-9F1E87/2020|EPI_ISL_589637|20 ···················· ···················· ····················· ···················· ···················· ····················
M 2640 hCoV-19/England/MILK-9F0D30/2020|EPI_ISL_589638|20 ···················· ···················· ····················· ···················· ···················· ····················
M 2641 hCoV-19/England/MILK-9F1A47/2020|EPI_ISL_589640|20 ···················· ···················· ····················· ···················· ···················· ····················
M 2642 hCoV-19/England/MILK-9F0E5B/2020|EPI_ISL_589641|20 ···················· ···················· ····················· ···················· ···················· ····················
M 2643 hCoV-19/England/MILK-9F186B/2020|EPI_ISL_589642|20 ···················· ···················· ····················· ···················· ···················· ····················
M 2644 hCoV-19/England/MILK-9F0ADF/2020|EPI_ISL_589643|20 ···················· ···················· ····················· ···················· ···················· ····················
M 2645 hCoV-19/England/MILK-9F1C14/2020|EPI_ISL_589645|20 ···················· ···················· ····················· ···················· ···················· ····················
M 2646 hCoV-19/Wales/MILK-9F1D5D/2020|EPI_ISL_589646|2020 ···················· ···················· ····················· ···················· ···················· ····················
M 2647 hCoV-19/Wales/MILK-9F1616/2020|EPI_ISL_589647|2020 ···················· ···················· ····················· ···················· ···················· ····················
M 2648 hCoV-19/England/MILK-9F0C33/2020|EPI_ISL_589648|20 ···················· ···················· ····················· ···················· ····T··············· ····················
M 2649 hCoV-19/England/MILK-9F19B3/2020|EPI_ISL_589649|20 ···················· ···················· ····················· ···················· ···················· ····················
M 2650 hCoV-19/England/MILK-9F0C24/2020|EPI_ISL_589651|20 ···················· ···················· ····················· ···················· ···················· ····················
M 2651 hCoV-19/England/MILK-9F0C8E/2020|EPI_ISL_589652|20 ···················· ···················· ····················· ···················· ···················· ····················
M 2652 hCoV-19/England/MILK-9F0E4C/2020|EPI_ISL_589653|20 ···················· ···················· ····················· ···················· ···················· ····················
M 2653 hCoV-19/England/MILK-9F190E/2020|EPI_ISL_589654|20 ···················· ·A·················· ····················· ···················· ···················· ···A················
M 2654 hCoV-19/England/MILK-9F1160/2020|EPI_ISL_589655|20 ···················· ···················· ····················· ···················· ···················· ····················
M 2655 hCoV-19/England/MILK-9F1115/2020|EPI_ISL_589656|20 ···················· ···················· ····················· ···················· ···················· ····················
M 2656 hCoV-19/England/QEUH-9B6F5E/2020|EPI_ISL_590600|20 ···················· ···················· ····················· ···················· ···················· ····················
M 2657 hCoV-19/England/QEUH-9B7791/2020|EPI_ISL_590601|20 ···················· ···················· ····················· ···················· ···················· ····················
M 2658 hCoV-19/England/QEUH-9B7E05/2020|EPI_ISL_590602|20 ···················· ···················· ····················· ···················· ···················· ····················
M 2659 hCoV-19/England/QEUH-9B70B4/2020|EPI_ISL_590603|20 ···················· ···················· ····················· ···················· ···················· ····················
M 2660 hCoV-19/England/QEUH-9B7AC5/2020|EPI_ISL_590604|20 ···················· ···················· ····················· ···················· ···················· ····················
M 2661 hCoV-19/England/QEUH-9B7E14/2020|EPI_ISL_590605|20 ···················· ·A·················· ····················· ···················· ···················· ···A················
M 2662 hCoV-19/England/QEUH-9B7F4E/2020|EPI_ISL_590606|20 ···················· ···················· ····················· ···················· ···················· ····················
```

| | | | | | | | | |
|---|---|---|---|---|---|---|---|---|
| M | 2663 hCoV-19/England/QEUH-9B76A3/2020\|EPI_ISL_590607\|20 | ···················· | ···················· | ···················· | ···················· | ···················· | ···················· | |
| M | 2664 hCoV-19/England/QEUH-9B72FA/2020\|EPI_ISL_590608\|20 | ···················· | ·A·················· | ···················· | ···················· | ···················· | ··A················· | |
| M | 2665 hCoV-19/England/QEUH-9B78E9/2020\|EPI_ISL_590609\|20 | ···················· | ···················· | ···················· | ···················· | ···················· | ···················· | |
| M | 2666 hCoV-19/England/QEUH-9B7843/2020\|EPI_ISL_590610\|20 | ···················· | ···················· | ···················· | ···················· | ···················· | ···················· | |
| M | 2667 hCoV-19/England/QEUH-9B7AD4/2020\|EPI_ISL_590611\|20 | ···················· | ···················· | ···················· | ···················· | ···················· | ···················· | |
| M | 2668 hCoV-19/England/QEUH-9B7D35/2020\|EPI_ISL_590612\|20 | ···················· | ···················· | ···················· | ···················· | ···················· | ···················· | |
| M | 2669 hCoV-19/England/QEUH-9B7360/2020\|EPI_ISL_590613\|20 | ···················· | ···················· | ···················· | ···················· | ···················· | ···················· | |
| M | 2670 hCoV-19/England/QEUH-9B77A0/2020\|EPI_ISL_590614\|20 | ···················· | ···················· | ···················· | ···················· | ···················· | ···················· | |
| M | 2671 hCoV-19/England/QEUH-9B7B95/2020\|EPI_ISL_590616\|20 | ···················· | ···A················ | ···················· | ···················· | ···················· | ···················· | |
| M | 2672 hCoV-19/England/QEUH-9B7148/2020\|EPI_ISL_590617\|20 | ···················· | ···················· | ···················· | ···················· | ···················· | ···················· | |
| M | 2673 hCoV-19/England/QEUH-9B7175/2020\|EPI_ISL_590618\|20 | ···················· | ···················· | ···················· | ···················· | ···················· | ···················· | |
| M | 2674 hCoV-19/England/QEUH-9B7324/2020\|EPI_ISL_590619\|20 | ···················· | ···················· | ···················· | ···················· | ···················· | ···················· | |
| M | 2675 hCoV-19/England/QEUH-9B72DC/2020\|EPI_ISL_590620\|20 | ···················· | ···················· | ···················· | ···················· | ···················· | ···················· | |
| M | 2676 hCoV-19/England/QEUH-9B7B3B/2020\|EPI_ISL_590621\|20 | ···················· | ···················· | ···················· | ···················· | ······T············· | ···················· | |
| M | 2677 hCoV-19/England/QEUH-9B705A/2020\|EPI_ISL_590622\|20 | ···················· | ···················· | ···················· | ···················· | ···················· | ···················· | |
| M | 2678 hCoV-19/England/QEUH-9B7B1D/2020\|EPI_ISL_590623\|20 | ···················· | ···················· | ···················· | ···················· | ···················· | ···················· | |
| M | 2679 hCoV-19/England/QEUH-9B800E/2020\|EPI_ISL_590624\|20 | ···················· | ···················· | ···················· | ···················· | ···················· | ···················· | |
| M | 2680 hCoV-19/England/QEUH-9B7E23/2020\|EPI_ISL_590625\|20 | ···················· | ···················· | ···················· | ···················· | ···················· | ···················· | |
| M | 2681 hCoV-19/England/QEUH-9B8174/2020\|EPI_ISL_590626\|20 | ···················· | ···················· | ···················· | ···················· | ···················· | ···················· | |
| M | 2682 hCoV-19/England/QEUH-9B6F30/2020\|EPI_ISL_590627\|20 | ···················· | ···················· | ···················· | ···················· | ···················· | ···················· | |
| M | 2683 hCoV-19/England/QEUH-9B7BC2/2020\|EPI_ISL_590629\|20 | ···················· | ···················· | ···················· | ···················· | ···················· | ···················· | |
| M | 2684 hCoV-19/England/QEUH-9B753D/2020\|EPI_ISL_590630\|20 | ···················· | ···················· | ···················· | ···················· | ···················· | ···················· | |
| M | 2685 hCoV-19/England/QEUH-9B79E6/2020\|EPI_ISL_590631\|20 | ···················· | ···················· | ···················· | ···················· | ···················· | ···················· | |
| M | 2686 hCoV-19/England/QEUH-9B7D71/2020\|EPI_ISL_590633\|20 | ···················· | ···················· | ···················· | ···················· | ···················· | ···················· | |
| M | 2687 hCoV-19/England/QEUH-9B799B/2020\|EPI_ISL_590634\|20 | ···················· | ···················· | ···················· | ···················· | ···················· | ···················· | |
| M | 2688 hCoV-19/England/QEUH-9B803B/2020\|EPI_ISL_590635\|20 | ···················· | ···················· | ···················· | ···················· | ···················· | ···················· | |
| M | 2689 hCoV-19/England/QEUH-9B702D/2020\|EPI_ISL_590636\|20 | ···················· | ···················· | ···················· | ···················· | ···················· | ···················· | |
| M | 2690 hCoV-19/England/QEUH-9B761C/2020\|EPI_ISL_590637\|20 | ···················· | ···················· | ···················· | ···················· | ···················· | ···················· | |
| M | 2691 hCoV-19/England/QEUH-9B81DE/2020\|EPI_ISL_590638\|20 | ···················· | ···················· | ···················· | ···················· | ···················· | ···················· | |
| M | 2692 hCoV-19/England/QEUH-9B7649/2020\|EPI_ISL_590639\|20 | ···················· | ···················· | ···················· | ···················· | ···················· | ···················· | |
| M | 2693 hCoV-19/Australia/VIC11943/2020\|EPI_ISL_591900\|202 | ···················· | ···················· | ···C················ | ···················· | ···················· | ···················· | |
| M | 2694 hCoV-19/Australia/VIC11950/2020\|EPI_ISL_591901\|202 | ···················· | ···················· | ···················· | ···················· | ···················· | ···················· | |
| M | 2695 hCoV-19/Australia/VIC11953/2020\|EPI_ISL_591902\|202 | ···················· | ···················· | ···K················ | ···················· | ···················· | ···················· | |
| M | 2696 hCoV-19/Australia/VIC11955/2020\|EPI_ISL_591903\|202 | ···················· | ···················· | ···K················ | ···················· | ···················· | ·R·················· | |
| M | 2697 hCoV-19/Australia/VIC11961/2020\|EPI_ISL_591904\|202 | ···················· | ···················· | ···················· | ···················· | ···················· | ···················· | |
| M | 2698 hCoV-19/Australia/VIC11983/2020\|EPI_ISL_591905\|202 | ···················· | ···················· | ···················· | ···················· | ···················· | ···················· | |
| M | 2699 hCoV-19/Australia/VIC11999/2020\|EPI_ISL_591908\|202 | ···················· | ···················· | ···················· | ···················· | ···················· | ···················· | |
| M | 2700 hCoV-19/Australia/VIC12033/2020\|EPI_ISL_591909\|202 | ···················· | ···················· | ···················· | ···················· | ···················· | ···················· | |
| M | 2701 hCoV-19/Australia/VIC12205/2020\|EPI_ISL_591930\|202 | ···················· | ···················· | ···················· | ···················· | ···················· | ···················· | |
| M | 2702 hCoV-19/Australia/VIC12211/2020\|EPI_ISL_591931\|202 | ···················· | ···················· | ···················· | ···················· | ···················· | ···················· | |
| M | 2703 hCoV-19/Australia/VIC12217/2020\|EPI_ISL_591932\|202 | ···················· | ···················· | ···················· | ···················· | ···················· | ···················· | |
| M | 2705 hCoV-19/Australia/VIC12225/2020\|EPI_ISL_591934\|202 | ···················· | ···················· | ···················· | ···················· | ···················· | ···················· | |
| M | 2706 hCoV-19/Australia/VIC12230/2020\|EPI_ISL_591935\|202 | ···················· | ···················· | ···················· | ···················· | ···················· | ···················· | |
| M | 2707 hCoV-19/Australia/VIC12254/2020\|EPI_ISL_591936\|202 | ···················· | ···················· | ···················· | ···················· | ···················· | ···················· | |
| M | 2709 hCoV-19/Australia/VIC12258/2020\|EPI_ISL_591938\|202 | ···················· | ···················· | ···················· | ···················· | ···················· | ···················· | |
| M | 2710 hCoV-19/Australia/VIC12267/2020\|EPI_ISL_591939\|202 | ···················· | ···················· | ···················· | ···················· | ···················· | ···················· | |
| M | 2711 hCoV-19/Australia/VIC12271/2020\|EPI_ISL_591940\|202 | ···················· | ···················· | ···················· | ···················· | ···················· | ···················· | |
| M | 2712 hCoV-19/Australia/VIC12272/2020\|EPI_ISL_591941\|202 | ···················· | ···················· | ···K················ | ···················· | ···················· | ···················· | |
| M | 2713 hCoV-19/Australia/VIC12273/2020\|EPI_ISL_591942\|202 | ···················· | ···················· | ···K················ | ···················· | ···················· | ···················· | |
| M | 2715 hCoV-19/Australia/VIC12275/2020\|EPI_ISL_591944\|202 | ···················· | ···················· | ···K················ | ···················· | ···················· | ···················· | |
| M | 2716 hCoV-19/Australia/VIC12276/2020\|EPI_ISL_591945\|202 | ···················· | ···················· | ···N················ | ···················· | ···················· | ·R·················· | |
| M | 2717 hCoV-19/Australia/VIC12277/2020\|EPI_ISL_591946\|202 | ···················· | ···················· | ···················· | ···················· | ···················· | ···················· | |
| M | 2718 hCoV-19/Australia/VIC12280/2020\|EPI_ISL_591947\|202 | ···················· | ···················· | ···················· | ···················· | ···················· | ···················· | |
| M | 2719 hCoV-19/Australia/VIC12281/2020\|EPI_ISL_591948\|202 | ···················· | ···················· | ···················· | ···················· | ···················· | ···················· | |
| M | 2720 hCoV-19/Australia/VIC12283/2020\|EPI_ISL_591949\|202 | ···················· | ···················· | ···K···K············ | ···················· | ···················· | ···················· | |
| M | 2721 hCoV-19/Australia/VIC12297/2020\|EPI_ISL_591950\|202 | ···················· | ···················· | ···K················ | ···················· | ···················· | ···················· | |
| M | 2722 hCoV-19/Australia/VIC12302/2020\|EPI_ISL_591951\|202 | ···················· | ···················· | ···K················ | ···················· | ···················· | ···················· | |
| M | 2723 hCoV-19/Australia/VIC12306/2020\|EPI_ISL_591952\|202 | ···················· | ···················· | ···················· | ···················· | ···················· | ···················· | |
| M | 2724 hCoV-19/Australia/VIC12307/2020\|EPI_ISL_591953\|202 | ···················· | ···················· | ···K················ | ···················· | ···················· | ···················· | |
| M | 2725 hCoV-19/Australia/VIC12310/2020\|EPI_ISL_591954\|202 | ···················· | ···················· | ···················· | ···················· | ···················· | ···················· | |
| M | 2726 hCoV-19/Australia/VIC12311/2020\|EPI_ISL_591955\|202 | ···················· | ···················· | ···················· | ···················· | ···················· | ···················· | |
| M | 2728 hCoV-19/Australia/VIC12319/2020\|EPI_ISL_591957\|202 | ···················· | ···················· | ···················· | ···················· | NNN················· | ···················· | 1 |
| M | 2729 hCoV-19/Australia/VIC12323/2020\|EPI_ISL_591958\|202 | ···················· | ···················· | ···N················ | ···················· | ···················· | ···················· | |
| M | 2730 hCoV-19/Australia/VIC12324/2020\|EPI_ISL_591959\|202 | ···················· | ···················· | ···K················ | ···················· | ···················· | ···················· | |
| M | 2731 hCoV-19/Australia/VIC12328/2020\|EPI_ISL_591960\|202 | ···················· | ···················· | ···K················ | ···················· | ···················· | ···················· | |
| M | 2732 hCoV-19/Australia/VIC12339/2020\|EPI_ISL_591961\|202 | ···················· | ···················· | ···················· | ···················· | NNN················· | ···················· | 1 |
| M | 2733 hCoV-19/Australia/VIC12341/2020\|EPI_ISL_591962\|202 | ···················· | ···················· | ···················· | ···················· | ···················· | ···················· | |
| M | 2735 hCoV-19/Australia/VIC12345/2020\|EPI_ISL_591964\|202 | ···················· | ···················· | ···················· | ···················· | ···················· | ···················· | |
| M | 2736 hCoV-19/Australia/VIC12348/2020\|EPI_ISL_591965\|202 | ···················· | ···················· | ···················· | ···················· | ···················· | ···················· | |
| M | 2737 hCoV-19/Australia/VIC12349/2020\|EPI_ISL_591966\|202 | ···················· | ···················· | ···················· | ···················· | ···················· | ···················· | |
| M | 2738 hCoV-19/Australia/VIC12367/2020\|EPI_ISL_591967\|202 | ···················· | ···················· | ···················· | ···················· | ···················· | ···················· | |
| M | 2739 hCoV-19/Australia/VIC12368/2020\|EPI_ISL_591969\|202 | ···················· | ···················· | ···················· | ···················· | ···················· | ···················· | |
| M | 2740 hCoV-19/USA/UT-UPHL-201007915/2020\|EPI_ISL_594101\| | ···················· | ···················· | ···················· | ···················· | ···················· | ···················· | |
| M | 2741 hCoV-19/USA/UT-UPHL-201017918/2020\|EPI_ISL_594102\| | ···················· | ···················· | ···················· | ···················· | ···················· | ···················· | |
| M | 2742 hCoV-19/USA/UT-UPHL-201009153/2020\|EPI_ISL_594103\| | ···················· | ···················· | ···················· | ···················· | ···················· | ···················· | |
| M | 2743 hCoV-19/USA/UT-UPHL-201008019/2020\|EPI_ISL_594104\| | ···················· | ···················· | ···················· | ···················· | ···················· | ···················· | |
| M | 2744 hCoV-19/USA/UT-UPHL-201017695/2020\|EPI_ISL_594106\| | ···················· | ···················· | ···················· | ···················· | ···················· | ···················· | |
| M | 2745 hCoV-19/USA/UT-UPHL-201022808/2020\|EPI_ISL_594107\| | ···················· | ···················· | ···················· | ···················· | ···················· | ···················· | |
| M | 2746 hCoV-19/USA/UT-UPHL-201021581/2020\|EPI_ISL_594110\| | ···················· | ···················· | ···················· | ···················· | ···················· | ···················· | |
| M | 2747 hCoV-19/USA/UT-UPHL-201014307/2020\|EPI_ISL_594111\| | ···················· | ···················· | ···················· | ···················· | ···················· | ···················· | |

| | | | | | | | | |
|---|---|---|---|---|---|---|---|---|
| M | 2748 hCoV-19/USA/UT-UPHL-201016815/2020\|EPI_ISL_594112\| | ···················· | ···················· | ···················· | ···················· | ···················· | ···················· | |
| M | 2749 hCoV-19/USA/UT-UPHL-201008588/2020\|EPI_ISL_594113\| | ···················· | ···················· | ···················· | ···················· | ···················· | ···················· | |
| M | 2750 hCoV-19/USA/UT-UPHL-201009968/2020\|EPI_ISL_594114\| | ···················· | ···················· | ···················· | ···················· | ···················· | ···················· | |
| M | 2751 hCoV-19/USA/UT-UPHL-201027414/2020\|EPI_ISL_594115\| | ···················· | ···················· | ···················· | ···················· | ···················· | ···················· | |
| M | 2752 hCoV-19/Ecuador/USFQ-1026/2020\|EPI_ISL_594118\|2020 | ···················· | ···················· | ···················· | ···················· | ···················· | ···················· | |
| M | 2753 hCoV-19/Sweden/20-52670/2020\|EPI_ISL_594148\|2020-0 | ···················· | ···················· | ···················· | ···················· | ···················· | ···················· | |
| M | 2754 hCoV-19/Sweden/20-08986/2020\|EPI_ISL_594149\|2020-0 | ···················· | ···················· | ···················· | ···················· | ···················· | ···················· | |
| M | 2755 hCoV-19/Sweden/20-08987/2020\|EPI_ISL_594150\|2020-0 | ···················· | ···················· | ···················· | ···················· | ···················· | ···················· | |
| M | 2756 hCoV-19/Sweden/20-08988/2020\|EPI_ISL_594151\|2020-0 | ···················· | ···················· | ···················· | ···················· | ···················· | ···················· | |
| M | 2757 hCoV-19/Sweden/20-08989/2020\|EPI_ISL_594152\|2020-0 | ···················· | ···················· | ···················· | ···················· | ···················· | ···················· | |
| M | 2758 hCoV-19/Sweden/20-08994/2020\|EPI_ISL_594153\|2020-0 | ···················· | ···················· | ···················· | ···················· | ···················· | ···················· | |
| M | 2759 hCoV-19/Sweden/20-51969/2020\|EPI_ISL_594154\|2020-0 | ···················· | ···················· | ···················· | ···················· | ···················· | ···················· | |
| M | 2760 hCoV-19/Israel/ISR_NH_MA_0727/2020\|EPI_ISL_594158\| | ···················· | ·A·················· | ···················· | ···················· | ······T············· | ···················· | |
| M | 2761 hCoV-19/Israel/ISR_NH_GD1_0714/2020\|EPI_ISL_594159 | ···················· | ···················· | ···················· | ···················· | ···················· | ···················· | |
| M | 2762 hCoV-19/Israel/ISR_NH_GD2_0714/2020\|EPI_ISL_594160 | ···················· | ···················· | ···················· | ···················· | ···················· | ···················· | |
| M | 2763 hCoV-19/Australia/WA267/2020\|EPI_ISL_594179\|2020-0 | ···················· | ···················· | ···················· | ···················· | ···················· | ···················· | |
| M | 2764 hCoV-19/Iran/2297/2020\|EPI_ISL_594185\|2020-04-13 | ···················· | ···················· | ···················· | ···················· | ···················· | ···················· | |
| M | 2765 hCoV-19/Iran/1325/2020\|EPI_ISL_594186\|2020-03-28 | ···················· | ···················· | ···················· | ···················· | ···················· | ···················· | |
| M | 2766 hCoV-19/Iran/2024/2020\|EPI_ISL_594187\|2020-04-11 | ···················· | ·····C·············· | ···················· | ···················· | ···················· | ···················· | |
| M | 2767 hCoV-19/Iran/7650/2020\|EPI_ISL_594188\|2020-06-26 | ···················· | ···················· | ···················· | ···················· | ···················· | ···················· | |
| M | 2768 hCoV-19/Wales/PHWC-47E124/2020\|EPI_ISL_595410\|2020 | ···················· | ···················· | ···················· | ···················· | ···················· | ···················· | |
| M | 2769 hCoV-19/Wales/PHWC-47E142/2020\|EPI_ISL_595412\|2020 | ···················· | ···················· | ···················· | ···················· | ···················· | ···················· | |
| M | 2770 hCoV-19/Wales/PHWC-47E151/2020\|EPI_ISL_595413\|2020 | ···················· | ···················· | ···················· | ···················· | ···················· | ···················· | |
| M | 2771 hCoV-19/Wales/PHWC-47E160/2020\|EPI_ISL_595414\|2020 | ···················· | ···················· | ···················· | ···················· | ···················· | ···················· | |
| M | 2772 hCoV-19/Wales/PHWC-47E18E/2020\|EPI_ISL_595416\|2020 | ···················· | ···················· | ···················· | ···················· | ···················· | ···················· | |
| M | 2773 hCoV-19/Wales/PHWC-47E19D/2020\|EPI_ISL_595417\|2020 | ···················· | ···················· | ···················· | ···················· | ···················· | ···················· | |
| M | 2775 hCoV-19/Wales/PHWC-47E203/2020\|EPI_ISL_595423\|2020 | ···················· | ·A·······A·········· | ···················· | ···················· | ···················· | ···················· | |
| M | 2776 hCoV-19/Wales/PHWC-47E212/2020\|EPI_ISL_595424\|2020 | ···················· | ···················· | ···················· | ···················· | ···················· | ···················· | |
| M | 2777 hCoV-19/Wales/PHWC-47E230/2020\|EPI_ISL_595426\|2020 | ···················· | ···················· | ···················· | ···················· | ···················· | ···················· | |
| M | 2778 hCoV-19/Wales/PHWC-47E24F/2020\|EPI_ISL_595427\|2020 | ···················· | ···················· | ···················· | ···················· | ···················· | ···················· | |
| M | 2779 hCoV-19/Wales/PHWC-47E25E/2020\|EPI_ISL_595428\|2020 | ···················· | ···················· | ···················· | ···················· | ···················· | ···················· | |
| M | 2780 hCoV-19/Wales/PHWC-47E27C/2020\|EPI_ISL_595429\|2020 | ···················· | ···················· | ···················· | ···················· | ···················· | ···················· | |
| M | 2781 hCoV-19/Wales/PHWC-47E28B/2020\|EPI_ISL_595430\|2020 | ···················· | ···················· | ···················· | ···················· | ···················· | ···················· | |
| M | 2782 hCoV-19/Wales/PHWC-47E2A9/2020\|EPI_ISL_595431\|2020 | ···················· | ···················· | ···················· | ···················· | ···················· | ···················· | |
| M | 2784 hCoV-19/Wales/PHWC-47E2D6/2020\|EPI_ISL_595434\|2020 | ···················· | ···················· | ···················· | ···················· | ···················· | ···················· | |
| M | 2785 hCoV-19/Wales/PHWC-47E2E5/2020\|EPI_ISL_595435\|2020 | ···················· | ···················· | ···················· | ···················· | ···················· | ···················· | |
| M | 2787 hCoV-19/Wales/PHWC-47E34C/2020\|EPI_ISL_595441\|2020 | ···················· | ···················· | ···················· | ···················· | ···················· | ···················· | |
| M | 2788 hCoV-19/Wales/PHWC-47E36A/2020\|EPI_ISL_595443\|2020 | ···················· | ···················· | ···················· | ···················· | ···················· | ···················· | |
| M | 2789 hCoV-19/Wales/PHWC-47E379/2020\|EPI_ISL_595444\|2020 | ···················· | ···················· | ···················· | ···················· | ···················· | ···················· | |
| M | 2790 hCoV-19/Wales/PHWC-47E388/2020\|EPI_ISL_595445\|2020 | ···················· | ···················· | ···················· | ···················· | ···················· | ···················· | |
| M | 2792 hCoV-19/Wales/PHWC-47E3A6/2020\|EPI_ISL_595447\|2020 | ···················· | ···················· | ···················· | ···················· | ···················· | ···················· | |
| M | 2793 hCoV-19/Wales/PHWC-47E3B5/2020\|EPI_ISL_595448\|2020 | ···················· | ···················· | ···················· | ···················· | ···················· | ···················· | |
| M | 2794 hCoV-19/Wales/PHWC-47E3C4/2020\|EPI_ISL_595449\|2020 | ···················· | ···················· | ···················· | ···················· | ···················· | ···················· | |
| M | 2795 hCoV-19/Wales/PHWC-47E3D3/2020\|EPI_ISL_595450\|2020 | ···················· | ···················· | ···················· | ···················· | ···················· | ···················· | |
| M | 2796 hCoV-19/Wales/PHWC-47E3E2/2020\|EPI_ISL_595451\|2020 | ···················· | ···················· | ···················· | ···················· | ···················· | ···················· | |
| M | 2797 hCoV-19/Wales/PHWC-47E3F1/2020\|EPI_ISL_595452\|2020 | ···················· | ···················· | ···················· | ···················· | ···················· | ···················· | |
| M | 2798 hCoV-19/Wales/PHWC-47E40D/2020\|EPI_ISL_595453\|2020 | ···················· | ···················· | ···················· | ···················· | ···················· | ···················· | |
| M | 2799 hCoV-19/Wales/PHWC-47E4EF/2020\|EPI_ISL_595463\|2020 | ···················· | ···················· | ···················· | ···················· | ···················· | ···················· | |
| M | 2800 hCoV-19/Wales/PHWC-47E4FE/2020\|EPI_ISL_595464\|2020 | ···················· | ···················· | ···················· | ···················· | ··A················· | ···················· | |
| M | 2801 hCoV-19/Wales/PHWC-47E5EC/2020\|EPI_ISL_595476\|2020 | ···················· | ···················· | ···················· | ···················· | ···················· | ···················· | |
| M | 2802 hCoV-19/Wales/PHWC-47E607/2020\|EPI_ISL_595478\|2020 | ···················· | ···················· | ···················· | ···················· | ···················· | ···················· | |
| M | 2803 hCoV-19/Wales/PHWC-47E661/2020\|EPI_ISL_595483\|2020 | ···················· | ···················· | ···················· | ···················· | ···················· | ···················· | |
| M | 2804 hCoV-19/Wales/PHWC-47E68F/2020\|EPI_ISL_595484\|2020 | ···················· | ···················· | ···················· | ···················· | ···················· | ···················· | |
| M | 2805 hCoV-19/Wales/PHWC-47E69E/2020\|EPI_ISL_595485\|2020 | ···················· | ···················· | ···················· | ···················· | ···················· | ···················· | |
| M | 2806 hCoV-19/Wales/PHWC-47E6AD/2020\|EPI_ISL_595486\|2020 | ···················· | ···················· | ···················· | ···················· | ·NNN················ | ···················· | 1 |
| M | 2807 hCoV-19/Wales/PHWC-47E6CB/2020\|EPI_ISL_595488\|2020 | ···················· | ···················· | ···················· | ···················· | ···················· | ···················· | |
| M | 2809 hCoV-19/Wales/PHWC-47E6E9/2020\|EPI_ISL_595490\|2020 | ···················· | ···················· | ···················· | ···················· | ···················· | ···················· | |
| M | 2810 hCoV-19/Wales/PHWC-47E6F8/2020\|EPI_ISL_595491\|2020 | ···················· | ···················· | ···················· | ···················· | ···················· | ···················· | |
| M | 2811 hCoV-19/Wales/PHWC-47E704/2020\|EPI_ISL_595492\|2020 | ···················· | ···················· | ···················· | ···················· | ···················· | ···················· | |
| M | 2812 hCoV-19/Wales/PHWC-47E713/2020\|EPI_ISL_595493\|2020 | ···················· | ···················· | ···················· | ···················· | ···················· | ···················· | |
| M | 2813 hCoV-19/Wales/PHWC-47E722/2020\|EPI_ISL_595494\|2020 | ···················· | ···················· | ···················· | ···················· | ···················· | ···················· | |
| M | 2814 hCoV-19/Wales/PHWC-47E740/2020\|EPI_ISL_595495\|2020 | ···················· | ···················· | ···················· | ···················· | ···················· | ···················· | |
| M | 2815 hCoV-19/Wales/PHWC-47E75F/2020\|EPI_ISL_595496\|2020 | ···················· | ···················· | ···················· | ···················· | ···················· | ···················· | |
| M | 2816 hCoV-19/Wales/PHWC-47E76E/2020\|EPI_ISL_595497\|2020 | ···················· | ···················· | ···················· | ···················· | ···················· | ···················· | |
| M | 2817 hCoV-19/Slovakia/UKBA-313/2020\|EPI_ISL_583481\|2020 | ···················· | ···················· | ···················· | ···················· | ···················· | ···················· | |
| M | 2818 hCoV-19/Slovakia/UKBA-314/2020\|EPI_ISL_583482\|2020 | ···················· | ···················· | ···················· | ···················· | ···················· | ···················· | |
| M | 2819 hCoV-19/Slovakia/UKBA-315/2020\|EPI_ISL_583483\|2020 | ···················· | ···················· | ···················· | ···················· | ···················· | ···················· | |
| M | 2820 hCoV-19/Slovakia/UKBA-316/2020\|EPI_ISL_583484\|2020 | ···················· | ···················· | ···················· | ···················· | ···················· | ···················· | |
| M | 2821 hCoV-19/Slovakia/UKBA-317/2020\|EPI_ISL_583485\|2020 | ···················· | ···················· | ···················· | ···················· | ···················· | ···················· | |
| M | 2822 hCoV-19/Slovakia/UKBA-318/2020\|EPI_ISL_583486\|2020 | ···················· | ···················· | ···················· | ···················· | ···················· | ···················· | |
| M | 2823 hCoV-19/Slovakia/UKBA-319/2020\|EPI_ISL_583487\|2020 | ···················· | ···················· | ···················· | ···················· | ···················· | ···················· | |
| M | 2824 hCoV-19/Slovakia/UKBA-321/2020\|EPI_ISL_583488\|2020 | ···················· | ···················· | ···················· | ···················· | ···················· | ···················· | |
| M | 2825 hCoV-19/Slovakia/UKBA-322/2020\|EPI_ISL_583489\|2020 | ···················· | ···················· | ···················· | ···················· | ···················· | ···················· | |
| M | 2826 hCoV-19/Brazil/SP-459/2020\|EPI_ISL_583490\|2020-04- | ···················· | ···················· | ···················· | ···················· | ···················· | ···················· | |
| M | 2827 hCoV-19/Brazil/SP-460/2020\|EPI_ISL_583491\|2020-04- | ···················· | ···················· | ···················· | ···················· | ···················· | ···················· | |
| M | 2828 hCoV-19/Brazil/SP-461/2020\|EPI_ISL_583492\|2020-04- | ···················· | ···················· | ···················· | ···················· | ···················· | ···················· | |
| M | 2829 hCoV-19/Brazil/SP-488/2020\|EPI_ISL_583493\|2020-04- | ···················· | ···················· | ···················· | ···················· | ···················· | ···················· | |
| M | 2830 hCoV-19/Brazil/SP-493/2020\|EPI_ISL_583494\|2020-04- | ···················· | ···················· | ···················· | ···················· | ···················· | ···················· | |
| M | 2831 hCoV-19/Brazil/SP-345/2020\|EPI_ISL_583495\|2020-06- | ···················· | ···················· | ···················· | ···················· | ···················· | ···················· | |
| M | 2832 hCoV-19/Brazil/SP-353/2020\|EPI_ISL_583496\|2020-06- | ···················· | ···················· | ···················· | ···················· | ···················· | ···················· | |

| | | | | | | | | |
|---|---|---|---|---|---|---|---|---|
| M | 2833 | hCoV-19/England/SHEF-CD306/2020\|EPI_ISL_585059\|202 | ···················· | ···················· | ····················· | ···················· | ···················· | ···················· |
| M | 2834 | hCoV-19/England/SHEF-CD62B/2020\|EPI_ISL_585060\|202 | ···················· | ···················· | ····················· | ···················· | ···················· | ···················· |
| M | 2835 | hCoV-19/England/SHEF-CD861/2020\|EPI_ISL_585061\|202 | ···················· | ···················· | ····················· | ···················· | ···················· | ···················· |
| M | 2836 | hCoV-19/England/SHEF-CDD71/2020\|EPI_ISL_585063\|202 | ···················· | ···················· | ····················· | ···················· | ···················· | ···················· |
| M | 2837 | hCoV-19/England/SHEF-CE01D/2020\|EPI_ISL_585064\|202 | ···················· | ···················· | ····················· | ···················· | ···················· | ···················· |
| M | 2838 | hCoV-19/England/SHEF-CE0B3/2020\|EPI_ISL_585065\|202 | ···················· | ···················· | ····················· | ···················· | ···················· | ···················· |
| M | 2839 | hCoV-19/England/SHEF-CE2EA/2020\|EPI_ISL_585066\|202 | ···················· | ···················· | ····················· | ···················· | ···················· | ···················· |
| M | 2840 | hCoV-19/England/SHEF-CE5A5/2020\|EPI_ISL_585068\|202 | ···················· | ···················· | ····················· | ···················· | ···················· | ···················· |
| M | 2841 | hCoV-19/England/SHEF-CE6A2/2020\|EPI_ISL_585069\|202 | ···················· | ···················· | ····················· | ···················· | ···················· | ···················· |
| M | 2842 | hCoV-19/England/SHEF-CEE40/2020\|EPI_ISL_585070\|202 | ···················· | ···················· | ····················· | ···················· | ···················· | ···················· |
| M | 2844 | hCoV-19/England/SHEF-CF119/2020\|EPI_ISL_585073\|202 | ···················· | ···················· | ····················· | ···················· | ···················· | ···················· |
| M | 2845 | hCoV-19/England/SHEF-CF28F/2020\|EPI_ISL_585074\|202 | ···················· | ···················· | ····················· | ···················· | ···················· | ···················· |
| M | 2846 | hCoV-19/England/SHEF-CFC09/2020\|EPI_ISL_585075\|202 | ···················· | ···················· | ····················· | ···················· | ···················· | ···················· |
| M | 2847 | hCoV-19/England/SHEF-D0E1E/2020\|EPI_ISL_585077\|202 | ···················· | ···················· | ····················· | ···················· | ···················· | ···················· |
| M | 2848 | hCoV-19/England/SHEF-D127B/2020\|EPI_ISL_585078\|202 | ···················· | ···················· | ····················· | ···················· | ···················· | ···················· |
| M | 2849 | hCoV-19/England/SHEF-D131E/2020\|EPI_ISL_585079\|202 | ···················· | ···················· | ····················· | ···················· | ···················· | ···················· |
| M | 2850 | hCoV-19/England/SHEF-D18F1/2020\|EPI_ISL_585081\|202 | ···················· | ···················· | ····················· | ···················· | ···················· | ···················· |
| M | 2851 | hCoV-19/England/SHEF-D233B/2020\|EPI_ISL_585084\|202 | ···················· | ···················· | ····················· | ···················· | ···················· | ···················· |
| M | 2854 | hCoV-19/England/SHEF-D33FE/2020\|EPI_ISL_585091\|202 | ···················· | ···················· | ····················· | ···················· | ···················· | ···················· |
| M | 2856 | hCoV-19/Northern Ireland/NIRE-106FA2/2020\|EPI_ISL_ | ···················· | ···················· | ····················· | ···················· | ···················· | ···················· |
| M | 2857 | hCoV-19/Northern Ireland/NIRE-106FC0/2020\|EPI_ISL_ | ···················· | ···················· | ····················· | ···················· | ···················· | ···················· |
| M | 2858 | hCoV-19/Northern Ireland/NIRE-106FDF/2020\|EPI_ISL_ | ···················· | ···················· | ····················· | ···················· | ···················· | ···················· |
| M | 2860 | hCoV-19/Northern Ireland/NIRE-106FFD/2020\|EPI_ISL_ | ···················· | ···················· | ····················· | ···················· | ···················· | ···················· |
| M | 2861 | hCoV-19/Northern Ireland/NIRE-107026/2020\|EPI_ISL_ | ···················· | ···················· | ····················· | ···················· | ···················· | ···················· |
| M | 2864 | hCoV-19/Northern Ireland/NIRE-10717E/2020\|EPI_ISL_ | ···················· | ···················· | ····················· | ···················· | ···················· | ···················· |
| M | 2865 | hCoV-19/Northern Ireland/NIRE-10718D/2020\|EPI_ISL_ | ···················· | ···················· | ····················· | ···················· | ···················· | ···················· |
| M | 2867 | hCoV-19/Northern Ireland/NIRE-1071AB/2020\|EPI_ISL_ | ···················· | ···················· | ····················· | ···················· | ···················· | ···················· |
| M | 2869 | hCoV-19/Northern Ireland/NIRE-1071C9/2020\|EPI_ISL_ | ···················· | ···················· | ····················· | ···················· | ···················· | ···················· |
| M | 2870 | hCoV-19/Northern Ireland/NIRE-1071D8/2020\|EPI_ISL_ | ···················· | ···················· | ····················· | ···················· | ···················· | ···················· |
| M | 2871 | hCoV-19/Northern Ireland/NIRE-1071E7/2020\|EPI_ISL_ | ···················· | ···················· | ····················· | ···················· | ···················· | ···················· |
| M | 2872 | hCoV-19/Canada/ON-S3/2020\|EPI_ISL_586390\|2020-03-1 | ···················· | ···················· | ····················· | ···················· | ···················· | ···················· |
| M | 2873 | hCoV-19/Canada/ON-S307/2020\|EPI_ISL_586391\|2020-03 | ···················· | ···················· | ····················· | ···················· | ···················· | ···················· |
| M | 2874 | hCoV-19/Canada/ON-S31/2020\|EPI_ISL_586393\|2020-03- | ···················· | ···················· | ····················· | ···················· | ···················· | ···················· |
| M | 2875 | hCoV-19/Canada/ON-S322/2020\|EPI_ISL_586396\|2020-03 | ···················· | ···················· | ····················· | ···················· | ···················· | ···················· |
| M | 2876 | hCoV-19/Canada/ON-S326/2020\|EPI_ISL_586398\|2020-03 | ···················· | ···················· | ····················· | ···················· | ···················· | ···················· |
| M | 2877 | hCoV-19/Canada/ON-S791/2020\|EPI_ISL_586446\|2020-04 | ···················· | ···················· | ····················· | ···················· | ···················· | ···················· |
| M | 2878 | hCoV-19/Canada/ON-S817/2020\|EPI_ISL_586450\|2020-04 | ···················· | ···················· | ····················· | ···················· | ···················· | ···················· |
| M | 2879 | hCoV-19/Canada/ON-S818/2020\|EPI_ISL_586451\|2020-04 | ···················· | ···················· | ····················· | ···················· | ·······Y············ | ···················· |
| M | 2880 | hCoV-19/Canada/ON-S824/2020\|EPI_ISL_586452\|2020-04 | ···················· | ···················· | ····················· | ···················· | ···················· | ···················· |
| M | 2881 | hCoV-19/Canada/ON-S831/2020\|EPI_ISL_586453\|2020-04 | ···················· | ···················· | ····················· | ···················· | ···················· | ···················· |
| M | 2882 | hCoV-19/Canada/ON-S834/2020\|EPI_ISL_586454\|2020-04 | ···················· | ···················· | ····················· | ···················· | ···················· | ···················· |
| M | 2883 | hCoV-19/Canada/ON-S835/2020\|EPI_ISL_586455\|2020-04 | ···················· | ···················· | ····················· | ···················· | ···················· | ···················· |
| M | 2884 | hCoV-19/England/ALDP-9FCE40/2020\|EPI_ISL_587776\|20 | ···················· | ···················· | ····················· | ···················· | ···················· | ···················· |
| M | 2885 | hCoV-19/England/ALDP-9FC772/2020\|EPI_ISL_587777\|20 | ···················· | ···················· | ····················· | ···················· | ···················· | ···················· |
| M | 2886 | hCoV-19/England/ALDP-9FCF5C/2020\|EPI_ISL_587779\|20 | ···················· | ···················· | ····················· | ···················· | ···················· | ···················· |
| M | 2887 | hCoV-19/England/ALDP-9FD261/2020\|EPI_ISL_587780\|20 | ···················· | ···················· | ····················· | ···················· | ···················· | ···················· |
| M | 2888 | hCoV-19/England/ALDP-9FD6DE/2020\|EPI_ISL_587781\|20 | ···················· | ···················· | ····················· | ···················· | ···················· | ···················· |
| M | 2889 | hCoV-19/England/ALDP-9FCAC4/2020\|EPI_ISL_587782\|20 | ···················· | ···················· | ····················· | ···················· | ···················· | ···················· |
| M | 2890 | hCoV-19/England/ALDP-9FCCBF/2020\|EPI_ISL_587783\|20 | ···················· | ···················· | ····················· | ···················· | ·····T·············· | ···················· |
| M | 2891 | hCoV-19/England/ALDP-9FD50E/2020\|EPI_ISL_587784\|20 | ···················· | ···················· | ····················· | ···················· | ···················· | ···················· |
| M | 2892 | hCoV-19/England/ALDP-9FD577/2020\|EPI_ISL_587785\|20 | ···················· | ···················· | ····················· | ···················· | ···················· | ···················· |
| M | 2893 | hCoV-19/England/ALDP-9FCE7D/2020\|EPI_ISL_587786\|20 | ···················· | ·A·················· | ····················· | ···················· | ···················· | ··A················· |
| M | 2894 | hCoV-19/England/ALDP-9FC9B8/2020\|EPI_ISL_587789\|20 | ···················· | ···················· | ····················· | ···················· | ···················· | ···················· |
| M | 2895 | hCoV-19/Australia/VIC13171/2020\|EPI_ISL_592200\|202 | ···················· | ···················· | ····················· | ···················· | ···················· | ···················· |
| M | 2896 | hCoV-19/Australia/VIC13172/2020\|EPI_ISL_592201\|202 | ···················· | ···················· | ····················· | ···················· | ···················· | ···················· |
| M | 2897 | hCoV-19/Australia/VIC13175/2020\|EPI_ISL_592202\|202 | ···················· | ···················· | ····················· | ···················· | ···················· | ···················· |
| M | 2898 | hCoV-19/Australia/VIC13177/2020\|EPI_ISL_592203\|202 | ···················· | ···················· | ····················· | ···················· | ···················· | ···················· |
| M | 2899 | hCoV-19/Australia/VIC13185/2020\|EPI_ISL_592204\|202 | ···················· | ···················· | ····················· | ···················· | ···················· | ···················· |
| M | 2900 | hCoV-19/Australia/VIC13191/2020\|EPI_ISL_592205\|202 | ···················· | ···················· | ····················· | ···················· | ···················· | ···················· |
| M | 2901 | hCoV-19/Australia/VIC13204/2020\|EPI_ISL_592206\|202 | ···················· | ···················· | ····················· | ···················· | ···················· | ···················· |
| M | 2902 | hCoV-19/Australia/VIC13206/2020\|EPI_ISL_592207\|202 | ···················· | ···················· | ····················· | ···················· | ···················· | ···················· |
| M | 2903 | hCoV-19/Australia/VIC13208/2020\|EPI_ISL_592208\|202 | ···················· | ···················· | ····················· | ···················· | ···················· | ···················· |
| M | 2904 | hCoV-19/Australia/VIC13209/2020\|EPI_ISL_592209\|202 | ···················· | ···················· | ····················· | ···················· | ···················· | ···················· |
| M | 2905 | hCoV-19/Australia/VIC13211/2020\|EPI_ISL_592210\|202 | ···················· | ···················· | ····················· | ···················· | ···················· | ···················· |
| M | 2906 | hCoV-19/Australia/VIC13212/2020\|EPI_ISL_592211\|202 | ···················· | ···················· | ····················· | ···················· | ···················· | ···················· |
| M | 2907 | hCoV-19/Australia/VIC13214/2020\|EPI_ISL_592212\|202 | ···················· | ···················· | ····················· | ···················· | ···················· | ···················· |
| M | 2908 | hCoV-19/Australia/VIC13217/2020\|EPI_ISL_592213\|202 | ···················· | ···················· | ····················· | ···················· | ···················· | ···················· |
| M | 2909 | hCoV-19/Australia/VIC13222/2020\|EPI_ISL_592214\|202 | ···················· | ···················· | ····················· | ···················· | ···················· | ···················· |
| M | 2910 | hCoV-19/Australia/VIC13229/2020\|EPI_ISL_592215\|202 | ···················· | ···················· | ····················· | ···················· | ···················· | ···················· |
| M | 2911 | hCoV-19/Australia/VIC13237/2020\|EPI_ISL_592216\|202 | ···················· | ···················· | ····················· | ···················· | ···················· | ···················· |
| M | 2912 | hCoV-19/Australia/VIC13246/2020\|EPI_ISL_592217\|202 | ···················· | ···················· | ····················· | ···················· | ···················· | ···················· |
| M | 2913 | hCoV-19/Australia/VIC13252/2020\|EPI_ISL_592218\|202 | ···················· | ···················· | ····················· | ···················· | ···················· | ···················· |
| M | 2914 | hCoV-19/Australia/VIC13257/2020\|EPI_ISL_592219\|202 | ···················· | ···················· | ····················· | ···················· | ···················· | ···················· |
| M | 2915 | hCoV-19/Australia/VIC13258/2020\|EPI_ISL_592220\|202 | ···················· | ···················· | ····················· | ···················· | ···················· | ···················· |
| M | 2917 | hCoV-19/Australia/VIC13273/2020\|EPI_ISL_592222\|202 | ···················· | ···················· | ····················· | ···················· | ···················· | ···················· |
| M | 2918 | hCoV-19/Australia/VIC13278/2020\|EPI_ISL_592223\|202 | ···················· | ···················· | ····················· | ···················· | ···················· | ···················· |
| M | 2919 | hCoV-19/Australia/VIC13288/2020\|EPI_ISL_592224\|202 | ···················· | ···················· | ····················· | ···················· | ···················· | ···················· |
| M | 2920 | hCoV-19/Australia/VIC13290/2020\|EPI_ISL_592225\|202 | ···················· | ···················· | ····················· | ···················· | ···················· | ···················· |
| M | 2921 | hCoV-19/Australia/VIC13310/2020\|EPI_ISL_592226\|202 | ···················· | ···················· | ····················· | ···················· | ···················· | ···················· |
| M | 2922 | hCoV-19/Australia/VIC13324/2020\|EPI_ISL_592227\|202 | ···················· | ···················· | ····················· | ···················· | ···················· | ···················· |

M 2923 hCoV-19/Australia/VIC13335/2020|EPI_ISL_592228|202 ···················· ···················· ···················· ···················· ···················· ····················
M 2924 hCoV-19/Australia/VIC13345/2020|EPI_ISL_592229|202 ···················· ···················· ···················· ···················· ···················· ····················
M 2925 hCoV-19/Australia/VIC13349/2020|EPI_ISL_592230|202 ···················· ···················· ···················· ···················· ···················· ····················
M 2926 hCoV-19/Australia/VIC13357/2020|EPI_ISL_592231|202 ···················· ···················· ···················· ···················· ···················· ····················
M 2927 hCoV-19/Australia/VIC13362/2020|EPI_ISL_592232|202 ···················· ···················· ···················· ···················· ···················· ····················
M 2928 hCoV-19/Australia/VIC13385/2020|EPI_ISL_592233|202 ···················· ···················· ···················· ···················· ···················· ····················
M 2929 hCoV-19/Australia/VIC13395/2020|EPI_ISL_592234|202 ···················· ···················· ···················· ···················· ···················· ····················
M 2931 hCoV-19/Australia/VIC13434/2020|EPI_ISL_592236|202 ···················· ···················· ···················· ···················· ···················· ····················
M 2932 hCoV-19/Australia/VIC13438/2020|EPI_ISL_592237|202 ···················· ···················· ···················· ···················· ···················· ····················
M 2933 hCoV-19/Australia/VIC13440/2020|EPI_ISL_592238|202 ···················· ···················· ···················· ···················· ···················· ····················
M 2934 hCoV-19/Australia/VIC13441/2020|EPI_ISL_592239|202 ···················· ···················· ···················· ···················· ···················· ····················
M 2935 hCoV-19/Australia/VIC13442/2020|EPI_ISL_592240|202 ···················· ···················· ···················· ···················· ···················· ····················
M 2936 hCoV-19/Australia/VIC13443/2020|EPI_ISL_592241|202 ···················· ···················· ···················· ···················· ···················· ····················
M 2937 hCoV-19/Australia/VIC13451/2020|EPI_ISL_592242|202 ···················· ···················· ···················· ···················· ···················· ····················
M 2938 hCoV-19/Australia/VIC13452/2020|EPI_ISL_592243|202 ···················· ···················· ···················· ···················· ···················· ····················
M 2939 hCoV-19/Australia/VIC13465/2020|EPI_ISL_592244|202 ···················· ···················· ···················· ···················· ···················· ····················
M 2940 hCoV-19/Australia/VIC13466/2020|EPI_ISL_592245|202 ···················· ···················· ···················· ···················· ···················· ····················
M 2941 hCoV-19/Australia/VIC13467/2020|EPI_ISL_592246|202 ···················· ···················· ···················· ···················· ···················· ····················
M 2942 hCoV-19/Australia/VIC13468/2020|EPI_ISL_592247|202 ···················· ···················· ···················· ···················· ···················· ····················
M 2943 hCoV-19/Australia/VIC13469/2020|EPI_ISL_592248|202 ···················· ···················· ···················· ···················· ···················· ····················
M 2944 hCoV-19/Australia/VIC13470/2020|EPI_ISL_592249|202 ···················· ···················· ···················· ···················· ···················· ····················
M 2945 hCoV-19/Australia/VIC13472/2020|EPI_ISL_592250|202 ···················· ···················· ···················· ···················· ···················· ····················
M 2946 hCoV-19/Australia/VIC13475/2020|EPI_ISL_592251|202 ···················· ···················· ···················· ···················· ···················· ····················
M 2947 hCoV-19/Australia/VIC13476/2020|EPI_ISL_592252|202 ···················· ···················· ···················· ···················· ···················· ····················
M 2948 hCoV-19/Australia/VIC13477/2020|EPI_ISL_592253|202 ···················· ···················· ···················· ···················· ···················· ····················
M 2949 hCoV-19/Australia/VIC13478/2020|EPI_ISL_592254|202 ···················· ···················· ···················· ···················· ···················· ····················
M 2950 hCoV-19/Australia/VIC13479/2020|EPI_ISL_592255|202 ···················· ···················· ···················· ···················· ···················· ····················
M 2951 hCoV-19/Australia/VIC13480/2020|EPI_ISL_592256|202 ···················· ···················· ···················· ···················· ···················· ····················
M 2952 hCoV-19/Australia/VIC13481/2020|EPI_ISL_592257|202 ···················· ···················· ···················· ···················· ···················· ····················
M 2953 hCoV-19/Australia/VIC13482/2020|EPI_ISL_592258|202 ···················· ···················· ···················· ···················· ···················· ····················
M 2954 hCoV-19/Australia/VIC13484/2020|EPI_ISL_592259|202 ···················· ···················· ···················· ···················· ···················· ····················
M 2955 hCoV-19/Australia/VIC13485/2020|EPI_ISL_592260|202 ···················· ···················· ···················· ···················· ···················· ····················
M 2956 hCoV-19/Australia/VIC13487/2020|EPI_ISL_592261|202 ···················· ···················· ···················· ···················· ···················· ····················
M 2957 hCoV-19/Australia/VIC13488/2020|EPI_ISL_592262|202 ···················· ···················· ···················· ···················· ···················· ····················
M 2958 hCoV-19/Australia/VIC13489/2020|EPI_ISL_592263|202 ···················· ···················· ···················· ···················· ···················· ····················
M 2959 hCoV-19/Canada/ON-E22/2020|EPI_ISL_593501|2020-05- ···················· ···················· ···················· ···················· ···················· ····················
M 2960 hCoV-19/Canada/ON-E24/2020|EPI_ISL_593502|2020-05- ···················· ···················· ···················· ···················· ···················· ····················
M 2961 hCoV-19/Canada/ON-E25/2020|EPI_ISL_593503|2020-05- ···················· ···················· ···················· ···················· ···················· ····················
M 2962 hCoV-19/Canada/ON-E26/2020|EPI_ISL_593504|2020-05- ···················· ···················· ···················· ···················· ···················· ····················
M 2963 hCoV-19/Canada/ON-E28/2020|EPI_ISL_593506|2020-05- ···················· ···················· ···················· ···················· ···················· ····················
M 2964 hCoV-19/Canada/ON-E29/2020|EPI_ISL_593507|2020-05- ···················· ···················· ···················· ···················· ···················· ····················
M 2965 hCoV-19/Canada/ON-E30/2020|EPI_ISL_593508|2020-05- ···················· ···················· ···················· ···················· ···················· ····················
M 2966 hCoV-19/Canada/ON-E31/2020|EPI_ISL_593509|2020-05- ···················· ···················· ···················· ···················· ···················· ····················
M 2967 hCoV-19/Canada/ON-E33/2020|EPI_ISL_593511|2020-05- ···················· ···················· ···················· ···················· ···················· ····················
M 2968 hCoV-19/Canada/ON-E34/2020|EPI_ISL_593512|2020-05- ···················· ···················· ···················· ···················· ···················· ····················
M 2969 hCoV-19/Canada/ON-E36/2020|EPI_ISL_593514|2020-05- ···················· ···················· ···················· ···················· ···················· ····················
M 2970 hCoV-19/Canada/ON-E37/2020|EPI_ISL_593515|2020-06- ···················· ···················· ···················· ···················· ···················· ····················
M 2971 hCoV-19/Canada/ON-E39/2020|EPI_ISL_593517|2020-06- ···················· ···················· ···················· ···················· ···················· ····················
M 2972 hCoV-19/Canada/ON-E40/2020|EPI_ISL_593518|2020-05- ···················· ···················· ···················· ···················· ···················· ····················
M 2973 hCoV-19/Canada/ON-E46/2020|EPI_ISL_593523|2020-05- ···················· ···················· ···················· ···················· ···················· ····················
M 2974 hCoV-19/Canada/ON-E47/2020|EPI_ISL_593524|2020-05- ···················· ···················· ···················· ···················· ···················· ····················
M 2975 hCoV-19/Canada/ON-E50/2020|EPI_ISL_593525|2020-05- ···················· ···················· ···················· ···················· ···················· ····················
M 2976 hCoV-19/Canada/ON-E51/2020|EPI_ISL_593526|2020-05- ···················· ···················· ···················· ···················· ···················· ····················
M 2977 hCoV-19/Canada/ON-E53/2020|EPI_ISL_593528|2020-05- ···················· ···················· ···················· ···················· ···················· ····················
M 2978 hCoV-19/Canada/ON-E55/2020|EPI_ISL_593529|2020-05- ···················· ···················· ···················· ···················· ···················· ····················
M 2979 hCoV-19/Canada/ON-E58/2020|EPI_ISL_593532|2020-06- ···················· ···················· ···················· ···················· ···················· ····················
M 2980 hCoV-19/Canada/ON-E62/2020|EPI_ISL_593536|2020-06- ···················· ···················· ···················· ···················· ···················· ····················
M 2981 hCoV-19/Canada/ON-E64/2020|EPI_ISL_593537|2020-06- ···················· ···················· ···················· ···················· ···················· ····················
M 2982 hCoV-19/Canada/ON-E65/2020|EPI_ISL_593538|2020-06- ···················· ···················· ···················· ···················· ···················· ····················
M 2983 hCoV-19/Canada/ON-E66/2020|EPI_ISL_593539|2020-06- ···················· ···················· ···················· ···················· ···················· ····················
M 2984 hCoV-19/Canada/ON-E67/2020|EPI_ISL_593540|2020-06- ···················· ···················· ···················· ···················· ···················· ····················
M 2985 hCoV-19/Canada/ON-E68/2020|EPI_ISL_593541|2020-06- ···················· ···················· ···················· ···················· ···················· ····················
M 2986 hCoV-19/Canada/ON-E70/2020|EPI_ISL_593543|2020-03- ···················· ···················· ···················· ···················· ···················· ····················
M 2987 hCoV-19/Canada/ON-E72/2020|EPI_ISL_593545|2020-04- ···················· ···················· ···················· ···················· ···················· ····················
M 2988 hCoV-19/Canada/ON-E73/2020|EPI_ISL_593546|2020-04- ···················· ···················· ···················· ···················· ···················· ····················
M 2989 hCoV-19/Canada/ON-E74/2020|EPI_ISL_593547|2020-04- ···················· ···················· ···················· ···················· ···················· ····················
M 2990 hCoV-19/Canada/ON-E75/2020|EPI_ISL_593548|2020-04- ···················· ···················· ···················· ···················· ···················· ····················
M 2991 hCoV-19/Canada/ON-E76/2020|EPI_ISL_593549|2020-05- ···················· ···················· ···················· ···················· ···················· ····················
M 2992 hCoV-19/Canada/ON-E77/2020|EPI_ISL_593550|2020-05- ···················· ···················· ···················· ···················· ···················· ····················
M 2993 hCoV-19/Canada/ON-E78/2020|EPI_ISL_593551|2020-05- ···················· ···················· ···················· ···················· ···················· ····················
M 2994 hCoV-19/USA/MA-JLL-D81/2020|EPI_ISL_593553|2020-06 ···················· ···················· ···················· ···················· ···················· ····················
M 2995 hCoV-19/USA/MA-JLL-D143/2020|EPI_ISL_593556|2020-0 ···················· ···················· ···················· ···················· ···················· ····················
M 2996 hCoV-19/USA/TX-DSHS-0829/2020|EPI_ISL_593572|2020- ···················· ···················· ···················· ···················· ···················· ····················
M 2997 hCoV-19/USA/TX-DSHS-0839/2020|EPI_ISL_593581|2020- ···················· ···················· ···················· ···················· ···················· ····················
M 2998 hCoV-19/Scotland/CVR5068/2020|EPI_ISL_594801|2020- ···················· ···················· ···················· ···················· ···················· ····················
M 2999 hCoV-19/Scotland/CVR5069/2020|EPI_ISL_594802|2020- ···················· ···················· ···················· ···················· N··················· NN·N················ 1
M 3000 hCoV-19/Scotland/CVR5072/2020|EPI_ISL_594805|2020- ···················· ···················· ···················· ···················· ···················· ····················
M 3001 hCoV-19/Scotland/CVR5073/2020|EPI_ISL_594806|2020- ···················· ···················· ···················· ···················· ···················· ····················
M 3002 hCoV-19/Scotland/CVR5074/2020|EPI_ISL_594807|2020- ···················· ···················· ···················· ···················· ···················· ····················
M 3003 hCoV-19/Scotland/CVR5076/2020|EPI_ISL_594809|2020- ···················· ···················· ···················· ···················· ···················· ····················

| | | | | | | | | | |
|---|---|---|---|---|---|---|---|---|---|
| M | 3004 | hCoV-19/Scotland/CVR5079/2020\|EPI_ISL_594812\|2020- | ···················· | ···················· | ···················· | ···················· | ···················· | ···················· | |
| M | 3005 | hCoV-19/Scotland/CVR5080/2020\|EPI_ISL_594813\|2020- | ···················· | ···················· | ···················· | ···················· | ···················· | ···················· | |
| M | 3006 | hCoV-19/Scotland/CVR5081/2020\|EPI_ISL_594814\|2020- | ···················· | ···················· | ···················· | ···················· | ···················· | ···················· | |
| M | 3007 | hCoV-19/Scotland/CVR5082/2020\|EPI_ISL_594815\|2020- | ···················· | ·A·················· | ···················· | ···················· | ···················· | ··A················· | |
| M | 3008 | hCoV-19/Scotland/EDB8033/2020\|EPI_ISL_594816\|2020- | ···················· | ···················· | ···················· | ···················· | ···················· | ···················· | |
| M | 3009 | hCoV-19/Scotland/EDB8035/2020\|EPI_ISL_594817\|2020- | ···················· | ···················· | ···················· | ···················· | ···················· | ···················· | |
| M | 3010 | hCoV-19/Scotland/EDB8043/2020\|EPI_ISL_594819\|2020- | ···················· | ···················· | ···················· | ···················· | ···················· | ···················· | |
| M | 3011 | hCoV-19/Scotland/EDB8044/2020\|EPI_ISL_594820\|2020- | ···················· | ···················· | ···················· | ···················· | ···················· | ···················· | |
| M | 3012 | hCoV-19/Scotland/EDB8045/2020\|EPI_ISL_594821\|2020- | ···················· | ···················· | ···················· | ···················· | ···················· | ···················· | |
| M | 3013 | hCoV-19/Scotland/EDB8046/2020\|EPI_ISL_594822\|2020- | ···················· | ···················· | ···················· | ···················· | ···················· | ···················· | |
| M | 3014 | hCoV-19/Scotland/EDB8048/2020\|EPI_ISL_594823\|2020- | ···················· | ···················· | ···················· | ···················· | ···················· | ···················· | |
| M | 3015 | hCoV-19/Scotland/EDB8049/2020\|EPI_ISL_594824\|2020- | ···················· | ···················· | ···················· | ···················· | ···················· | ···················· | |
| M | 3016 | hCoV-19/Scotland/EDB8050/2020\|EPI_ISL_594825\|2020- | ···················· | ···················· | ···················· | ···················· | ···················· | ···················· | |
| M | 3018 | hCoV-19/Scotland/EDB8114/2020\|EPI_ISL_594829\|2020- | ···················· | ···················· | ···················· | ···················· | ···················· | ···················· | |
| M | 3019 | hCoV-19/Scotland/EDB8116/2020\|EPI_ISL_594830\|2020- | ···················· | ···················· | ···················· | ···················· | ···················· | ···················· | |
| M | 3020 | hCoV-19/Scotland/EDB8159/2020\|EPI_ISL_594832\|2020- | ···················· | ···················· | ···················· | ···················· | ···················· | ···················· | |
| M | 3021 | hCoV-19/Scotland/EDB8205/2020\|EPI_ISL_594833\|2020- | ···················· | ···················· | ···················· | ···················· | ···················· | ···················· | |
| M | 3022 | hCoV-19/Scotland/EDB8211/2020\|EPI_ISL_594834\|2020- | ···················· | ···················· | ···················· | ···················· | ···················· | ···················· | |
| M | 3023 | hCoV-19/Scotland/EDB8224/2020\|EPI_ISL_594835\|2020- | ···················· | ···················· | ···················· | ···················· | ···················· | ···················· | |
| M | 3025 | hCoV-19/Scotland/EDB8284/2020\|EPI_ISL_594838\|2020- | ···················· | ···················· | ···················· | ···················· | ···················· | ···················· | |
| M | 3026 | hCoV-19/Scotland/EDB8288/2020\|EPI_ISL_594839\|2020- | ···················· | ···················· | ···················· | ···················· | ···················· | ···················· | |
| M | 3027 | hCoV-19/Scotland/EDB8289/2020\|EPI_ISL_594840\|2020- | ···················· | ···················· | ···················· | ···················· | ···················· | ···················· | |
| M | 3030 | hCoV-19/Scotland/EDB8304/2020\|EPI_ISL_594844\|2020- | ···················· | ···················· | ···················· | ···················· | ···················· | ···················· | |
| M | 3032 | hCoV-19/England/LOND-12624ED/2020\|EPI_ISL_594861\|2 | ···················· | ···················· | ···················· | ···················· | ···················· | ···················· | |
| M | 3033 | hCoV-19/England/LOND-126259F/2020\|EPI_ISL_594862\|2 | ···················· | ·A·················· | ···················· | ···················· | ···················· | ··A················· | |
| M | 3034 | hCoV-19/England/LOND-12625AE/2020\|EPI_ISL_594863\|2 | ···················· | ···················· | ···················· | ···················· | ···················· | ···················· | |
| M | 3035 | hCoV-19/England/LOND-12625BD/2020\|EPI_ISL_594864\|2 | ···················· | ·A·················· | ···················· | ···················· | ···················· | ··A················· | |
| M | 3036 | hCoV-19/England/LOND-12625DB/2020\|EPI_ISL_594865\|2 | ···················· | ·A·················· | ···················· | ···················· | ···T················ | ··A················· | |
| M | 3037 | hCoV-19/England/LOND-1262632/2020\|EPI_ISL_594867\|2 | ···················· | ···················· | ···················· | ···················· | ···················· | ···················· | |
| M | 3038 | hCoV-19/England/LOND-1262650/2020\|EPI_ISL_594869\|2 | ···················· | ···················· | ···················· | ···················· | ···················· | ···················· | |
| M | 3039 | hCoV-19/England/LOND-12626C9/2020\|EPI_ISL_594870\|2 | ···················· | ···················· | ···················· | ···················· | ···················· | ···················· | |
| M | 3040 | hCoV-19/England/LOND-1262711/2020\|EPI_ISL_594871\|2 | ···················· | ···················· | ···················· | ···················· | ···················· | ···················· | |
| M | 3041 | hCoV-19/England/LOND-126275D/2020\|EPI_ISL_594872\|2 | ···················· | ···················· | ···················· | ···················· | ···················· | ···················· | |
| M | 3042 | hCoV-19/England/LOND-12627B7/2020\|EPI_ISL_594873\|2 | ···················· | ·A·················· | ···················· | ···················· | ···················· | ··A················· | |
| M | 3043 | hCoV-19/England/LOND-12627D5/2020\|EPI_ISL_594874\|2 | ···················· | ·A·················· | ···················· | ···················· | ···················· | ··A················· | |
| M | 3044 | hCoV-19/England/LOND-12627E4/2020\|EPI_ISL_594875\|2 | ···················· | ···················· | ···················· | ···················· | ···················· | ···················· | |
| M | 3045 | hCoV-19/England/LOND-1262869/2020\|EPI_ISL_594876\|2 | ···················· | ·A·················· | ···················· | ···················· | ···················· | ··A················· | |
| M | 3046 | hCoV-19/England/LOND-1262896/2020\|EPI_ISL_594878\|2 | ···················· | ···················· | ···················· | ···················· | ···················· | ···················· | |
| M | 3047 | hCoV-19/England/LOND-12628C3/2020\|EPI_ISL_594879\|2 | ···················· | ···················· | ···················· | ···················· | ···················· | ···················· | |
| M | 3048 | hCoV-19/England/LOND-126291B/2020\|EPI_ISL_594881\|2 | ···················· | ·A·················· | ···················· | ···················· | ···················· | ··A················· | |
| M | 3049 | hCoV-19/England/LOND-126292A/2020\|EPI_ISL_594882\|2 | ···················· | ···················· | ···················· | ···················· | ···················· | ···················· | |
| M | 3050 | hCoV-19/England/LOND-1262A36/2020\|EPI_ISL_594884\|2 | ···················· | ···················· | ···················· | ···················· | ···················· | ···················· | |
| M | 3051 | hCoV-19/England/LOND-1262A72/2020\|EPI_ISL_594885\|2 | ···················· | ···················· | ···················· | ···················· | ···················· | ···················· | |
| M | 3052 | hCoV-19/England/LOND-1262ACD/2020\|EPI_ISL_594886\|2 | ···················· | ···················· | ···················· | ···················· | ···················· | ···················· | |
| M | 3053 | hCoV-19/England/LOND-1262B33/2020\|EPI_ISL_594887\|2 | ···················· | ···················· | ···················· | ···················· | ···················· | ···················· | |
| M | 3054 | hCoV-19/England/MILK-9EEF6C/2020\|EPI_ISL_594890\|20 | ···················· | ·A·················· | ···················· | ···················· | ···················· | ··A················· | |
| M | 3055 | hCoV-19/Wales/MILK-9EEFA8/2020\|EPI_ISL_594891\|2020 | ···················· | ···················· | ···················· | ···················· | ···················· | ···················· | |
| M | 3056 | hCoV-19/England/MILK-9EEFC6/2020\|EPI_ISL_594892\|20 | ···················· | ·A·················· | ···················· | ···················· | ···················· | ··A················· | |
| M | 3057 | hCoV-19/Wales/MILK-9EF00E/2020\|EPI_ISL_594893\|2020 | ···················· | ···················· | ···················· | ···················· | ···················· | ···················· | |
| M | 3058 | hCoV-19/England/MILK-9EF04A/2020\|EPI_ISL_594895\|20 | ···················· | ···················· | ···········T········ | ···················· | ····T··············· | ···················· | |
| M | 3059 | hCoV-19/England/MILK-9EF068/2020\|EPI_ISL_594896\|20 | ···················· | ···················· | ···················· | ···················· | ···················· | ···················· | |
| M | 3060 | hCoV-19/England/MILK-9EF086/2020\|EPI_ISL_594897\|20 | ···················· | ···················· | ···················· | ···················· | ···················· | ···················· | |
| M | 3061 | hCoV-19/Wales/MILK-9EF095/2020\|EPI_ISL_594898\|2020 | ···················· | ···················· | ···················· | ···················· | ······T············· | ···················· | |
| M | 3062 | hCoV-19/Wales/MILK-9EF0D1/2020\|EPI_ISL_594899\|2020 | ···················· | ···················· | ···················· | ···················· | ···················· | ···················· | |
| M | 3064 | hCoV-19/Wales/PHWC-47E43A/2020\|EPI_ISL_595455\|2020 | ···················· | ···················· | ···················· | ···················· | ···················· | ···················· | |
| M | 3065 | hCoV-19/Wales/PHWC-47E467/2020\|EPI_ISL_595457\|2020 | ···················· | ···················· | ···················· | ···················· | ···················· | ···················· | |
| M | 3066 | hCoV-19/Wales/PHWC-47E4A3/2020\|EPI_ISL_595459\|2020 | ···················· | ···················· | ···················· | ···················· | ···················· | ···················· | |
| M | 3067 | hCoV-19/Wales/PHWC-47E50A/2020\|EPI_ISL_595465\|2020 | ···················· | ···················· | ···················· | ···················· | ···················· | ···················· | |
| M | 3068 | hCoV-19/Wales/PHWC-47E546/2020\|EPI_ISL_595467\|2020 | ···················· | ···················· | ···················· | ···················· | ···················· | ···················· | |
| M | 3069 | hCoV-19/Wales/PHWC-47E582/2020\|EPI_ISL_595470\|2020 | ···················· | ···················· | ···················· | ···················· | ···················· | ···················· | |
| M | 3070 | hCoV-19/Wales/PHWC-47E591/2020\|EPI_ISL_595471\|2020 | ···················· | ···················· | ···················· | ···················· | ···················· | ···················· | |
| M | 3071 | hCoV-19/Wales/PHWC-47E5BF/2020\|EPI_ISL_595473\|2020 | ···················· | ···················· | ···················· | ···················· | ···················· | ···················· | |
| M | 3072 | hCoV-19/Wales/PHWC-47E5DD/2020\|EPI_ISL_595475\|2020 | ···················· | ···················· | ···················· | ···················· | ···················· | ···················· | |
| M | 3075 | hCoV-19/Northern Ireland/NIRE-107071/2020\|EPI_ISL_ | ···················· | ···················· | ···················· | ···················· | ···················· | ···················· | |
| M | 3078 | hCoV-19/Northern Ireland/NIRE-1070BD/2020\|EPI_ISL_ | ···················· | ···················· | ···················· | ···················· | ···················· | ···················· | |
| M | 3079 | hCoV-19/Northern Ireland/NIRE-1070CC/2020\|EPI_ISL_ | ···················· | ···················· | ···················· | ···················· | ·NNN················ | ···················· | 1 |
| M | 3080 | hCoV-19/Northern Ireland/NIRE-1070DB/2020\|EPI_ISL_ | ···················· | ···················· | ···················· | ···················· | ···················· | ···················· | |
| M | 3081 | hCoV-19/Northern Ireland/NIRE-1070EA/2020\|EPI_ISL_ | ···················· | ···················· | ···················· | ···················· | ···················· | ···················· | |
| M | 3083 | hCoV-19/Northern Ireland/NIRE-107105/2020\|EPI_ISL_ | ···················· | ···················· | ···················· | ···················· | ···················· | ···················· | |
| M | 3085 | hCoV-19/Northern Ireland/NIRE-107211/2020\|EPI_ISL_ | ···················· | ···················· | ···················· | ···················· | ···················· | ···················· | |
| M | 3086 | hCoV-19/Northern Ireland/NIRE-107220/2020\|EPI_ISL_ | ···················· | ·············A······ | ···················· | ···················· | ···················· | ···················· | |
| M | 3087 | hCoV-19/Northern Ireland/NIRE-10723F/2020\|EPI_ISL_ | ···················· | ···················· | ···················· | ···················· | ···················· | ···················· | |
| M | 3090 | hCoV-19/Northern Ireland/NIRE-10728A/2020\|EPI_ISL_ | ···················· | ···················· | ···················· | ···················· | ···················· | ···················· | |
| M | 3091 | hCoV-19/Northern Ireland/NIRE-107299/2020\|EPI_ISL_ | ···················· | ···················· | ···················· | ···················· | ···NNN·············· | ···················· | 1 |
| M | 3093 | hCoV-19/Northern Ireland/NIRE-10730F/2020\|EPI_ISL_ | ···················· | ···················· | ···················· | ···················· | ···················· | ···················· | |
| M | 3094 | hCoV-19/Northern Ireland/NIRE-10731E/2020\|EPI_ISL_ | ···················· | ···················· | ···················· | ···················· | ···················· | ···················· | |
| M | 3096 | hCoV-19/Northern Ireland/NIRE-10733C/2020\|EPI_ISL_ | ···················· | ···················· | ···················· | ···················· | ···················· | ···················· | |
| M | 3097 | hCoV-19/Northern Ireland/NIRE-10734B/2020\|EPI_ISL_ | ···················· | ···················· | ···················· | ···················· | ···················· | ···················· | |
| M | 3098 | hCoV-19/Northern Ireland/NIRE-107387/2020\|EPI_ISL_ | ···················· | ···················· | ···················· | ···················· | ···················· | ···················· | |
| M | 3099 | hCoV-19/Northern Ireland/NIRE-1073A5/2020\|EPI_ISL_ | ···················· | ···················· | ···················· | ···················· | ···················· | ···················· | |

| | | | | | | | | |
|---|---|---|---|---|---|---|---|---|
| M | 3100 | hCoV-19/Northern Ireland/NIRE-1073C3/2020\|EPI_ISL_ | .................... | .................... | .................... | .................... | .................... | .................... |
| M | 3101 | hCoV-19/Northern Ireland/NIRE-1073E1/2020\|EPI_ISL_ | .................... | .................... | .................... | .................... | .................... | .................... |
| M | 3102 | hCoV-19/Northern Ireland/NIRE-1073F0/2020\|EPI_ISL_ | .................... | .................... | .................... | .................... | .................... | .................... |
| M | 3103 | hCoV-19/Northern Ireland/NIRE-10740C/2020\|EPI_ISL_ | .................... | .................... | .................... | .................... | .................... | .................... |
| M | 3104 | hCoV-19/Northern Ireland/NIRE-10741B/2020\|EPI_ISL_ | .................... | .................... | .................... | .................... | .................... | .................... |
| M | 3106 | hCoV-19/Northern Ireland/NIRE-107448/2020\|EPI_ISL_ | .................... | .................... | .................... | .................... | .................... | .................... |
| M | 3107 | hCoV-19/Northern Ireland/NIRE-107457/2020\|EPI_ISL_ | .................... | .................... | .................... | .................... | .................... | .................... |
| M | 3108 | hCoV-19/Northern Ireland/NIRE-107466/2020\|EPI_ISL_ | .................... | .................... | .................... | .................... | .................... | .................... |
| M | 3109 | hCoV-19/Northern Ireland/NIRE-107475/2020\|EPI_ISL_ | .................... | .................... | .................... | .................... | .................... | .................... |
| M | 3111 | hCoV-19/Northern Ireland/NIRE-107493/2020\|EPI_ISL_ | .................... | .................... | .................... | .................... | .................... | .................... |
| M | 3113 | hCoV-19/Northern Ireland/NIRE-1074B1/2020\|EPI_ISL_ | .................... | .................... | .................... | .................... | .................... | .................... |
| M | 3115 | hCoV-19/Northern Ireland/NIRE-1074DF/2020\|EPI_ISL_ | .................... | .................... | .................... | .................... | .................... | .................... |
| M | 3116 | hCoV-19/Northern Ireland/NIRE-1074EE/2020\|EPI_ISL_ | .................... | .................... | .................... | .................... | .................... | .................... |
| M | 3118 | hCoV-19/Northern Ireland/NIRE-107509/2020\|EPI_ISL_ | .................... | .................... | .................... | .................... | .................... | .................... |
| M | 3119 | hCoV-19/Northern Ireland/NIRE-107518/2020\|EPI_ISL_ | .................... | .................... | .................... | .................... | .................... | .................... |
| M | 3120 | hCoV-19/Northern Ireland/NIRE-107527/2020\|EPI_ISL_ | .................... | .................... | .................... | .................... | .................... | .................... |
| M | 3121 | hCoV-19/Northern Ireland/NIRE-107536/2020\|EPI_ISL_ | .................... | .................... | .................... | .................... | .................... | .................... |
| M | 3122 | hCoV-19/Northern Ireland/NIRE-107545/2020\|EPI_ISL_ | .................... | .................... | .................... | .................... | .................... | .................... |
| M | 3123 | hCoV-19/Northern Ireland/NIRE-107563/2020\|EPI_ISL_ | .................... | .................... | .................... | .................... | .................... | .................... |
| M | 3124 | hCoV-19/Northern Ireland/NIRE-107572/2020\|EPI_ISL_ | .................... | .................... | .................... | .................... | .................... | .................... |
| M | 3125 | hCoV-19/Northern Ireland/NIRE-107581/2020\|EPI_ISL_ | .................... | .................... | .................... | .................... | .................... | .................... |
| M | 3126 | hCoV-19/Northern Ireland/NIRE-107590/2020\|EPI_ISL_ | .................... | .................... | .................... | .................... | .................... | .................... |
| M | 3127 | hCoV-19/Northern Ireland/NIRE-1075AF/2020\|EPI_ISL_ | .................... | .................... | .................... | .................... | .................... | .................... |
| M | 3128 | hCoV-19/Northern Ireland/NIRE-1075BE/2020\|EPI_ISL_ | .................... | .................... | .................... | .................... | .................... | .................... |
| M | 3129 | hCoV-19/Northern Ireland/NIRE-1075CD/2020\|EPI_ISL_ | .................... | .................... | .................... | .................... | .................... | .................... |
| M | 3130 | hCoV-19/Northern Ireland/NIRE-108210/2020\|EPI_ISL_ | .................... | .................... | .................... | .................... | .................... | .................... |
| M | 3131 | hCoV-19/Northern Ireland/NIRE-10822F/2020\|EPI_ISL_ | .................... | .................... | .................... | .................... | .................... | .................... |
| M | 3135 | hCoV-19/Canada/ON-S357/2020\|EPI_ISL_586405\|2020-03 | .................... | .................... | .................... | .................... | .................... | .................... |
| M | 3136 | hCoV-19/Canada/ON-S46/2020\|EPI_ISL_586411\|2020-03- | .................... | .................... | .................... | .................... | .................... | .................... |
| M | 3137 | hCoV-19/Canada/ON-S719/2020\|EPI_ISL_586429\|2020-04 | .................... | .................... | .................... | .................... | .................... | .................... |
| M | 3138 | hCoV-19/Canada/ON-S738/2020\|EPI_ISL_586431\|2020-04 | .................... | .................... | .................... | .................... | .................... | .................... |
| M | 3139 | hCoV-19/Canada/ON-S739/2020\|EPI_ISL_586432\|2020-04 | .................... | .................... | .................... | .................... | .................... | .................... |
| M | 3140 | hCoV-19/Canada/ON-S741/2020\|EPI_ISL_586434\|2020-04 | .................... | .................... | .................... | .................... | .................... | .................... |
| M | 3141 | hCoV-19/Canada/ON-S749/2020\|EPI_ISL_586435\|2020-04 | .................... | .................... | .................... | .................... | ........Y........... | .................... |
| M | 3142 | hCoV-19/Canada/ON-S750/2020\|EPI_ISL_586437\|2020-04 | .................... | .................... | .................... | .................... | .................... | .................... |
| M | 3143 | hCoV-19/Canada/ON-S757/2020\|EPI_ISL_586438\|2020-04 | .................... | .................... | .................... | .................... | .................... | .................... |
| M | 3144 | hCoV-19/Canada/ON-S763/2020\|EPI_ISL_586439\|2020-04 | .................... | .................... | .................... | .................... | .................... | .................... |
| M | 3145 | hCoV-19/Canada/ON-S781/2020\|EPI_ISL_586442\|2020-04 | .................... | .................... | .................... | .................... | .................... | .................... |
| M | 3146 | hCoV-19/Canada/ON-S782/2020\|EPI_ISL_586443\|2020-04 | .................... | .................... | .................... | .................... | .................... | .................... |
| M | 3147 | hCoV-19/Canada/ON-S784/2020\|EPI_ISL_586444\|2020-04 | .................... | .................... | .................... | .................... | .................... | .................... |
| M | 3148 | hCoV-19/Canada/ON-S842/2020\|EPI_ISL_586457\|2020-04 | .................... | .................... | .................... | .................... | .................... | .................... |
| M | 3149 | hCoV-19/Canada/ON-S854/2020\|EPI_ISL_586459\|2020-04 | .................... | .................... | .................... | .................... | .................... | .................... |
| M | 3150 | hCoV-19/Canada/ON-S855/2020\|EPI_ISL_586460\|2020-04 | .................... | .................... | .................... | .................... | .................... | .................... |
| M | 3151 | hCoV-19/Canada/ON-S856/2020\|EPI_ISL_586461\|2020-04 | .................... | .................... | .................... | .................... | .................... | .................... |
| M | 3152 | hCoV-19/Canada/ON-S860/2020\|EPI_ISL_586462\|2020-04 | .................... | .................... | .................... | .................... | .................... | .................... |
| M | 3153 | hCoV-19/Canada/ON-S861/2020\|EPI_ISL_586463\|2020-04 | .................... | .................... | .................... | .................... | .................... | .................... |
| M | 3154 | hCoV-19/Canada/ON-S864/2020\|EPI_ISL_586464\|2020-04 | .................... | .................... | .................... | .................... | .................... | .................... |
| M | 3155 | hCoV-19/Canada/ON-S867/2020\|EPI_ISL_586465\|2020-04 | .................... | .................... | .................... | .................... | .................... | .................... |
| M | 3156 | hCoV-19/Canada/ON-S877/2020\|EPI_ISL_586467\|2020-04 | .................... | .................... | .................... | .................... | .................... | .................... |
| M | 3157 | hCoV-19/Canada/ON-S882/2020\|EPI_ISL_586469\|2020-04 | .................... | .................... | .................... | .................... | .................... | .................... |
| M | 3158 | hCoV-19/Canada/ON-S889/2020\|EPI_ISL_586471\|2020-04 | .................... | .................... | .................... | .................... | .................... | .................... |
| M | 3159 | hCoV-19/Canada/ON-S898/2020\|EPI_ISL_586473\|2020-04 | .................... | .................... | .................... | .................... | .................... | .................... |
| M | 3160 | hCoV-19/Canada/ON-S901/2020\|EPI_ISL_586474\|2020-04 | .................... | .................... | .................... | .................... | .................... | .................... |
| M | 3161 | hCoV-19/Canada/ON-S902/2020\|EPI_ISL_586475\|2020-04 | .................... | .................... | .................... | .................... | .................... | .................... |
| M | 3162 | hCoV-19/Canada/ON-S903/2020\|EPI_ISL_586476\|2020-04 | .................... | .................... | .................... | .................... | .................... | .................... |
| M | 3163 | hCoV-19/Canada/ON-S904/2020\|EPI_ISL_586477\|2020-04 | .................... | .................... | .................... | .................... | .................... | .................... |
| M | 3164 | hCoV-19/Canada/ON-S905/2020\|EPI_ISL_586478\|2020-04 | .................... | .................... | .................... | .................... | .................... | .................... |
| M | 3165 | hCoV-19/Canada/ON-S906/2020\|EPI_ISL_586479\|2020-04 | .................... | .................... | .................... | .................... | .................... | .................... |
| M | 3166 | hCoV-19/Canada/ON-S907/2020\|EPI_ISL_586480\|2020-04 | .................... | .................... | .................... | .................... | .................... | .................... |
| M | 3167 | hCoV-19/Canada/ON-S911/2020\|EPI_ISL_586482\|2020-04 | .................... | .................... | .................... | .................... | .................... | .................... |
| M | 3168 | hCoV-19/Canada/ON-S915/2020\|EPI_ISL_586483\|2020-04 | .................... | .................... | .................... | .................... | .................... | ...A................ |
| M | 3169 | hCoV-19/Canada/ON-S916/2020\|EPI_ISL_586484\|2020-04 | .................... | .................... | .................... | .................... | .................... | .................... |
| M | 3170 | hCoV-19/Canada/ON-S917/2020\|EPI_ISL_586485\|2020-04 | .................... | .................... | .................... | .................... | .................... | .................... |
| M | 3171 | hCoV-19/Canada/ON-S918/2020\|EPI_ISL_586486\|2020-04 | .................... | .................... | .................... | .................... | .................... | .................... |
| M | 3172 | hCoV-19/Canada/ON-S919/2020\|EPI_ISL_586487\|2020-04 | .................... | .................... | .................... | .................... | .................... | .................... |
| M | 3173 | hCoV-19/Canada/ON-S920/2020\|EPI_ISL_586488\|2020-04 | .................... | .................... | .................... | .................... | .................... | .................... |
| M | 3174 | hCoV-19/Canada/ON-S932/2020\|EPI_ISL_586490\|2020-04 | .................... | .................... | .................... | .................... | .................... | .................... |
| M | 3175 | hCoV-19/Canada/ON-S933/2020\|EPI_ISL_586491\|2020-04 | .................... | .................... | .................... | .................... | .................... | .................... |
| M | 3176 | hCoV-19/Canada/ON-S934/2020\|EPI_ISL_586492\|2020-04 | .................... | .................... | .................... | .................... | .................... | .................... |
| M | 3177 | hCoV-19/Canada/ON-S935/2020\|EPI_ISL_586493\|2020-04 | .................... | .................... | .................... | .................... | .................... | .................... |
| M | 3178 | hCoV-19/Canada/ON-S936/2020\|EPI_ISL_586494\|2020-04 | .................... | .................... | .................... | .................... | .................... | .................... |
| M | 3179 | hCoV-19/Canada/ON-S938/2020\|EPI_ISL_586495\|2020-04 | .................... | .................... | .................... | .................... | .................... | .................... |
| M | 3180 | hCoV-19/Canada/ON-S945/2020\|EPI_ISL_586498\|2020-04 | .................... | .................... | .................... | .................... | .................... | .................... |
| M | 3181 | hCoV-19/Canada/ON-S946/2020\|EPI_ISL_586499\|2020-04 | .................... | .................... | .................... | .................... | .................... | .................... |
| M | 3182 | hCoV-19/England/QEUH-A210E1/2020\|EPI_ISL_587700\|20 | .................... | .................... | .................... | .................... | .................... | .................... |
| M | 3183 | hCoV-19/England/QEUH-A20774/2020\|EPI_ISL_587701\|20 | .................... | .................... | .................... | .................... | .................... | .................... |
| M | 3184 | hCoV-19/England/QEUH-A2247B/2020\|EPI_ISL_587702\|20 | .................... | .................... | .................... | .................... | .................... | .................... |
| M | 3185 | hCoV-19/England/QEUH-A20325/2020\|EPI_ISL_587703\|20 | .................... | .................... | .................... | .................... | .................... | .................... |
| M | 3186 | hCoV-19/England/QEUH-A2071A/2020\|EPI_ISL_587704\|20 | .................... | .................... | .................... | .................... | .................... | .................... |
| M | 3187 | hCoV-19/England/QEUH-A22402/2020\|EPI_ISL_587705\|20 | .................... | .................... | .................... | .................... | .................... | .................... |

| | | | | | | | | |
|---|---|---|---|---|---|---|---|---|
| M | 3188 | hCoV-19/England/QEUH-A21E14/2020\|EPI_ISL_587706\|20 | ···················· | ···················· | ···················· | ···················· | ···················· | ···················· |
| M | 3189 | hCoV-19/England/QEUH-A20738/2020\|EPI_ISL_587709\|20 | ···················· | ···················· | ···················· | ···················· | ···················· | ···················· |
| M | 3190 | hCoV-19/England/QEUH-A20343/2020\|EPI_ISL_587710\|20 | ···················· | ···················· | ···················· | ···················· | ···················· | ···················· |
| M | 3191 | hCoV-19/England/ALDP-9FCE31/2020\|EPI_ISL_587711\|20 | ···················· | ···················· | ···················· | ···················· | ···················· | ···················· |
| M | 3192 | hCoV-19/England/ALDP-9FC8D9/2020\|EPI_ISL_587712\|20 | ···················· | ···················· | ···················· | ···················· | ···················· | ···················· |
| M | 3193 | hCoV-19/England/ALDP-9FDB75/2020\|EPI_ISL_587713\|20 | ···················· | ···················· | ···················· | ···················· | ···················· | ···················· |
| M | 3194 | hCoV-19/England/ALDP-9FD5FF/2020\|EPI_ISL_587714\|20 | ···················· | ···················· | ···················· | ···················· | ···················· | ···················· |
| M | 3195 | hCoV-19/England/ALDP-9FD3B9/2020\|EPI_ISL_587715\|20 | ···················· | ···················· | ···················· | ···················· | ···················· | ···················· |
| M | 3196 | hCoV-19/England/ALDP-9FD0D0/2020\|EPI_ISL_587716\|20 | ···················· | ···················· | ···················· | ···················· | ···················· | ···················· |
| M | 3197 | hCoV-19/England/ALDP-9FD5B3/2020\|EPI_ISL_587718\|20 | ···················· | ···················· | ···················· | ···················· | ···················· | ···················· |
| M | 3198 | hCoV-19/England/ALDP-9FD94E/2020\|EPI_ISL_587719\|20 | ···················· | ···················· | ···················· | ···················· | ···················· | ···················· |
| M | 3199 | hCoV-19/England/ALDP-9FD7EA/2020\|EPI_ISL_587720\|20 | ···················· | ···················· | ···················· | ···················· | ···················· | ···················· |
| M | 3200 | hCoV-19/England/ALDP-9FD60B/2020\|EPI_ISL_587722\|20 | ···················· | ···················· | ···················· | ···················· | ···················· | ···················· |
| M | 3201 | hCoV-19/England/ALDP-9FD8F6/2020\|EPI_ISL_587723\|20 | ············Y······· | ···················· | ···················· | ···················· | ···················· | ···················· |
| M | 3202 | hCoV-19/England/ALDP-9FDD8E/2020\|EPI_ISL_587724\|20 | ···················· | ···················· | ···················· | ···················· | ···················· | ···················· |
| M | 3203 | hCoV-19/England/ALDP-9FDCDC/2020\|EPI_ISL_587725\|20 | ···················· | ···················· | ···················· | ···················· | ···················· | ···················· |
| M | 3204 | hCoV-19/England/ALDP-9FCA6A/2020\|EPI_ISL_587726\|20 | ···················· | ···················· | ···················· | ···················· | ···················· | ···················· |
| M | 3205 | hCoV-19/England/ALDP-9FCEF5/2020\|EPI_ISL_587727\|20 | ···················· | ···················· | ···················· | ···················· | ···················· | ···················· |
| M | 3206 | hCoV-19/England/ALDP-9FC89D/2020\|EPI_ISL_587729\|20 | ···················· | ···················· | ···················· | ···················· | ···················· | ···················· |
| M | 3207 | hCoV-19/England/ALDP-9FDA69/2020\|EPI_ISL_587730\|20 | ···················· | ···················· | ···················· | ···················· | ···················· | ···················· |
| M | 3208 | hCoV-19/England/ALDP-9FCF7A/2020\|EPI_ISL_587731\|20 | ···················· | ···················· | ···················· | ···················· | ···················· | ···················· |
| M | 3209 | hCoV-19/England/ALDP-9FD191/2020\|EPI_ISL_587732\|20 | ···················· | ···················· | ···················· | ···················· | ·····T·············· | ···················· |
| M | 3210 | hCoV-19/England/ALDP-9FD322/2020\|EPI_ISL_587733\|20 | ···················· | ···················· | ···················· | ···················· | ···················· | ···················· |
| M | 3211 | hCoV-19/England/ALDP-9FD638/2020\|EPI_ISL_587734\|20 | ···················· | ···················· | ···················· | ···················· | ···················· | ···················· |
| M | 3212 | hCoV-19/England/ALDP-9FD8D8/2020\|EPI_ISL_587735\|20 | ···················· | ·A·················· | ···················· | ···················· | ···················· | ···················· |
| M | 3213 | hCoV-19/England/ALDP-9FD43E/2020\|EPI_ISL_587736\|20 | ···················· | ···················· | ···················· | ···················· | ···················· | ···················· |
| M | 3214 | hCoV-19/England/ALDP-9FD805/2020\|EPI_ISL_587737\|20 | ···················· | ·A·················· | ···················· | ···················· | ···················· | ···················· |
| M | 3215 | hCoV-19/England/ALDP-9FCB49/2020\|EPI_ISL_587738\|20 | ···················· | ···················· | ···················· | ···················· | ···················· | ···················· |
| M | 3216 | hCoV-19/England/ALDP-9FD234/2020\|EPI_ISL_587739\|20 | ···················· | ···················· | ···················· | ···················· | ···················· | ···················· |
| M | 3217 | hCoV-19/England/ALDP-9FD8E7/2020\|EPI_ISL_587740\|20 | ···················· | ···················· | ···················· | ···················· | ···················· | ···················· |
| M | 3218 | hCoV-19/England/ALDP-9FDE6D/2020\|EPI_ISL_587741\|20 | ···················· | ···················· | ···················· | ···················· | ···················· | ···················· |
| M | 3219 | hCoV-19/England/ALDP-9FCD70/2020\|EPI_ISL_587742\|20 | ···················· | ···················· | ···················· | ···················· | ···················· | ···················· |
| M | 3220 | hCoV-19/England/ALDP-9FDC54/2020\|EPI_ISL_587743\|20 | ···················· | ···················· | ···················· | ···················· | ···················· | ···················· |
| M | 3221 | hCoV-19/England/ALDP-9FD771/2020\|EPI_ISL_587744\|20 | ···················· | ·A·················· | ···················· | ···················· | ···················· | ··A················· |
| M | 3222 | hCoV-19/England/ALDP-9FD8C9/2020\|EPI_ISL_587745\|20 | ···················· | ···················· | ···················· | ···················· | ···················· | ···················· |
| M | 3223 | hCoV-19/England/ALDP-9FCED7/2020\|EPI_ISL_587746\|20 | ···················· | ···················· | ···················· | ···················· | ···················· | ···················· |
| M | 3224 | hCoV-19/England/ALDP-9FD28F/2020\|EPI_ISL_587747\|20 | ···················· | ···················· | ···················· | ···················· | ···················· | ···················· |
| M | 3225 | hCoV-19/England/ALDP-9FDB39/2020\|EPI_ISL_587748\|20 | ···················· | ···················· | ···················· | ···················· | ···················· | ···················· |
| M | 3226 | hCoV-19/England/ALDP-9FD058/2020\|EPI_ISL_587749\|20 | ···················· | ···················· | ···················· | ···················· | ···················· | ···················· |
| M | 3227 | hCoV-19/England/ALDP-9FD7BD/2020\|EPI_ISL_587750\|20 | ···················· | ·A·················· | ···················· | ···················· | ···················· | ··A················· |
| M | 3228 | hCoV-19/England/ALDP-9FD146/2020\|EPI_ISL_587752\|20 | ···················· | ···················· | ···················· | ···················· | ···················· | ···················· |
| M | 3229 | hCoV-19/England/ALDP-9FCEAA/2020\|EPI_ISL_587753\|20 | ···················· | ···················· | ···················· | ···················· | ···················· | ···················· |
| M | 3230 | hCoV-19/England/ALDP-9FD1FB/2020\|EPI_ISL_587756\|20 | ···················· | ···················· | ···················· | ···················· | ···················· | ···················· |
| M | 3231 | hCoV-19/England/ALDP-9FD243/2020\|EPI_ISL_587757\|20 | ···················· | ···················· | ···················· | ···················· | ···················· | ···················· |
| M | 3232 | hCoV-19/England/ALDP-9FDAC3/2020\|EPI_ISL_587758\|20 | ···················· | ···················· | ···················· | ···················· | ···················· | ···················· |
| M | 3233 | hCoV-19/England/ALDP-9FE00C/2020\|EPI_ISL_587760\|20 | ···················· | ···················· | ···················· | ···················· | ···················· | ···················· |
| M | 3234 | hCoV-19/England/ALDP-9FDA78/2020\|EPI_ISL_587761\|20 | ···················· | ···················· | ···················· | ···················· | ···················· | ···················· |
| M | 3235 | hCoV-19/England/ALDP-9FDA4B/2020\|EPI_ISL_587762\|20 | ···················· | ···················· | ···················· | ···················· | ···················· | ···················· |
| M | 3236 | hCoV-19/England/ALDP-9FC87F/2020\|EPI_ISL_587763\|20 | ···················· | ···················· | ···················· | ···················· | ···················· | ···················· |
| M | 3237 | hCoV-19/England/ALDP-9FD735/2020\|EPI_ISL_587764\|20 | ···················· | ···················· | ···················· | ···················· | ···················· | ···················· |
| M | 3238 | hCoV-19/England/ALDP-9FCC28/2020\|EPI_ISL_587765\|20 | ···················· | ···················· | ···················· | ···················· | ···················· | ···················· |
| M | 3239 | hCoV-19/England/ALDP-9FDD7F/2020\|EPI_ISL_587766\|20 | ···················· | ···················· | ···················· | ···················· | ···················· | ···················· |
| M | 3240 | hCoV-19/England/ALDP-9FD98A/2020\|EPI_ISL_587767\|20 | ···················· | ·A·················· | ···················· | ···················· | ···················· | ··A················· |
| M | 3241 | hCoV-19/England/ALDP-9FCBB2/2020\|EPI_ISL_587768\|20 | ···················· | ···················· | ···················· | ···················· | ···················· | ···················· |
| M | 3242 | hCoV-19/England/ALDP-9FCDCB/2020\|EPI_ISL_587769\|20 | ···················· | ···················· | ···················· | ···················· | ···················· | ···················· |
| M | 3243 | hCoV-19/England/ALDP-9FCBFE/2020\|EPI_ISL_587770\|20 | ···················· | ···················· | ···················· | ···················· | ···················· | ···················· |
| M | 3244 | hCoV-19/England/ALDP-9FD88D/2020\|EPI_ISL_587771\|20 | ···················· | ···················· | ···················· | ···················· | ···················· | ···················· |
| M | 3245 | hCoV-19/England/ALDP-9FC912/2020\|EPI_ISL_587772\|20 | ···················· | ···················· | ···················· | ···················· | ···················· | ···················· |
| M | 3246 | hCoV-19/England/ALDP-9FCE5F/2020\|EPI_ISL_587773\|20 | ···················· | ···················· | ···················· | ···················· | ···················· | ···················· |
| M | 3247 | hCoV-19/England/ALDP-9FD744/2020\|EPI_ISL_587774\|20 | ···················· | ···················· | ···················· | ···················· | ···················· | ···················· |
| M | 3248 | hCoV-19/England/ALDP-9FC930/2020\|EPI_ISL_587775\|20 | ···················· | ···················· | ···················· | ···················· | ···················· | ···················· |
| M | 3249 | hCoV-19/England/ALDP-9FDE8B/2020\|EPI_ISL_587787\|20 | ···················· | ···················· | ···················· | ···················· | ···················· | ···················· |
| M | 3250 | hCoV-19/England/ALDP-9FD61A/2020\|EPI_ISL_587788\|20 | ···················· | ···················· | ···················· | ···················· | ···················· | ···················· |
| M | 3251 | hCoV-19/England/ALDP-9FCA79/2020\|EPI_ISL_587791\|20 | ···················· | ···················· | ···················· | ···················· | ···················· | ···················· |
| M | 3252 | hCoV-19/England/ALDP-9FCFD4/2020\|EPI_ISL_587792\|20 | ···················· | ···················· | ···················· | ···················· | ···················· | ···················· |
| M | 3253 | hCoV-19/England/ALDP-9FD4D4/2020\|EPI_ISL_587794\|20 | ···················· | ···················· | ···················· | ···················· | ···················· | ···················· |
| M | 3254 | hCoV-19/England/ALDP-9FD049/2020\|EPI_ISL_587798\|20 | ···················· | ···················· | ···················· | ···················· | ···················· | ···················· |
| M | 3255 | hCoV-19/England/ALDP-9FDEC7/2020\|EPI_ISL_587799\|20 | ···················· | ···················· | ···················· | ···················· | ···················· | ···················· |
| M | 3256 | hCoV-19/Australia/VIC13491/2020\|EPI_ISL_592264\|202 | ···················· | ···················· | ···················· | ···················· | ···················· | ···················· |
| M | 3257 | hCoV-19/Australia/VIC13492/2020\|EPI_ISL_592265\|202 | ···················· | ···················· | ···················· | ···················· | ···················· | ···················· |
| M | 3258 | hCoV-19/Australia/VIC13493/2020\|EPI_ISL_592266\|202 | ···················· | ···················· | ···················· | ···················· | ···················· | ···················· |
| M | 3259 | hCoV-19/Australia/VIC13494/2020\|EPI_ISL_592267\|202 | ···················· | ···················· | ···················· | ···················· | ···················· | ···················· |
| M | 3260 | hCoV-19/Australia/VIC13496/2020\|EPI_ISL_592268\|202 | ···················· | ···················· | ···················· | ···················· | ···················· | ···················· |
| M | 3261 | hCoV-19/Australia/VIC13497/2020\|EPI_ISL_592269\|202 | ···················· | ···················· | ···················· | ···················· | ···················· | ···················· |
| M | 3262 | hCoV-19/Australia/VIC13499/2020\|EPI_ISL_592270\|202 | ···················· | ···················· | ···················· | ···················· | ···················· | ···················· |
| M | 3263 | hCoV-19/Australia/VIC13500/2020\|EPI_ISL_592271\|202 | ···················· | ···················· | ···················· | ···················· | ···················· | ···················· |
| M | 3264 | hCoV-19/Australia/VIC13501/2020\|EPI_ISL_592272\|202 | ···················· | ···················· | ···················· | ···················· | ···················· | ···················· |
| M | 3265 | hCoV-19/Australia/VIC13502/2020\|EPI_ISL_592273\|202 | ···················· | ···················· | ···················· | ···················· | ···················· | ···················· |
| M | 3266 | hCoV-19/Australia/VIC13503/2020\|EPI_ISL_592274\|202 | ···················· | ···················· | ···················· | ···················· | ···················· | ···················· |
| M | 3267 | hCoV-19/Australia/VIC13504/2020\|EPI_ISL_592275\|202 | ···················· | ···················· | ···················· | ···················· | ···················· | ···················· |

| | | | | | | | | |
|---|---|---|---|---|---|---|---|---|
| M | 3268 hCoV-19/Australia/VIC13505/2020\|EPI_ISL_592276\|202 | ···················· | ······A············· | ···················· | ···················· | ···················· | ···················· | |
| M | 3269 hCoV-19/Australia/VIC13506/2020\|EPI_ISL_592277\|202 | ···················· | ···················· | ···················· | ···················· | ···················· | ···················· | |
| M | 3270 hCoV-19/Australia/VIC13509/2020\|EPI_ISL_592278\|202 | ···················· | ···················· | ···················· | ···················· | ···················· | ···················· | |
| M | 3271 hCoV-19/Australia/VIC13510/2020\|EPI_ISL_592279\|202 | ···················· | ···················· | ···················· | ···················· | ···················· | ···················· | |
| M | 3272 hCoV-19/Australia/VIC13511/2020\|EPI_ISL_592280\|202 | ···················· | ···················· | ···················· | ···················· | ···················· | ···················· | |
| M | 3273 hCoV-19/Australia/VIC13513/2020\|EPI_ISL_592281\|202 | ···················· | ···················· | ···················· | ···················· | ···················· | ···················· | |
| M | 3274 hCoV-19/Australia/VIC13517/2020\|EPI_ISL_592282\|202 | ···················· | ···················· | ···················· | ···················· | ···················· | ···················· | |
| M | 3275 hCoV-19/Australia/VIC13519/2020\|EPI_ISL_592283\|202 | ···················· | ···················· | ···················· | ···················· | ···················· | ···················· | |
| M | 3276 hCoV-19/Australia/VIC13522/2020\|EPI_ISL_592284\|202 | ···················· | ···················· | ···················· | ···················· | ···················· | ···················· | |
| M | 3277 hCoV-19/Australia/VIC13523/2020\|EPI_ISL_592285\|202 | ···················· | ···················· | ···················· | ···················· | ···················· | ···················· | |
| M | 3278 hCoV-19/Australia/VIC13524/2020\|EPI_ISL_592286\|202 | ···················· | ···················· | ···················· | ···················· | ···················· | ···················· | |
| M | 3279 hCoV-19/Australia/VIC13525/2020\|EPI_ISL_592287\|202 | ···················· | ···················· | ···················· | ···················· | ···················· | ···················· | |
| M | 3280 hCoV-19/Australia/VIC13528/2020\|EPI_ISL_592288\|202 | ···················· | ···················· | ···················· | ···················· | ···················· | ···················· | |
| M | 3281 hCoV-19/Australia/VIC13532/2020\|EPI_ISL_592289\|202 | ···················· | ···················· | ···················· | ···················· | ···················· | ···················· | |
| M | 3282 hCoV-19/Australia/VIC13614/2020\|EPI_ISL_592290\|202 | ···················· | ···················· | ···················· | ···················· | ···················· | ···················· | |
| M | 3283 hCoV-19/Australia/VIC13618/2020\|EPI_ISL_592291\|202 | ···················· | ···················· | ···················· | ···················· | ···················· | ···················· | |
| M | 3284 hCoV-19/Australia/VIC13629/2020\|EPI_ISL_592292\|202 | ···················· | ···················· | ···················· | ···················· | ···················· | ···················· | |
| M | 3285 hCoV-19/Australia/VIC13633/2020\|EPI_ISL_592293\|202 | ···················· | ···················· | ···················· | ···················· | ···················· | ···················· | |
| M | 3286 hCoV-19/Australia/VIC13637/2020\|EPI_ISL_592294\|202 | ···················· | ···················· | ···················· | ···················· | ···················· | ···················· | |
| M | 3287 hCoV-19/Australia/VIC13683/2020\|EPI_ISL_592295\|202 | ···················· | ···················· | ···················· | ···················· | ···················· | ···················· | |
| M | 3288 hCoV-19/Australia/VIC13684/2020\|EPI_ISL_592296\|202 | ···················· | ···················· | ···················· | ···················· | ···················· | ···················· | |
| M | 3289 hCoV-19/Australia/VIC13687/2020\|EPI_ISL_592297\|202 | ···················· | ···················· | ···················· | ···················· | ···················· | ···················· | |
| M | 3290 hCoV-19/Australia/VIC13689/2020\|EPI_ISL_592298\|202 | ···················· | ···················· | ···················· | ···················· | ···················· | ···················· | |
| M | 3291 hCoV-19/Australia/VIC13690/2020\|EPI_ISL_592299\|202 | ···················· | ···················· | ···················· | ···················· | ···················· | ···················· | |
| M | 3292 hCoV-19/Australia/VIC15675/2020\|EPI_ISL_593410\|202 | ···················· | ···················· | ···················· | ···················· | ···················· | ···················· | |
| M | 3293 hCoV-19/Australia/VIC15683/2020\|EPI_ISL_593411\|202 | ···················· | ···················· | ···················· | ···················· | ···················· | ···················· | |
| M | 3294 hCoV-19/Australia/VIC15685/2020\|EPI_ISL_593412\|202 | ···················· | ···················· | ···················· | ···················· | ···················· | ···················· | |
| M | 3295 hCoV-19/Australia/VIC15698/2020\|EPI_ISL_593413\|202 | ···················· | ···················· | ···················· | ···················· | ···················· | ···················· | |
| M | 3296 hCoV-19/Australia/VIC15700/2020\|EPI_ISL_593414\|202 | ···················· | ···················· | ···················· | ···················· | ···················· | ···················· | |
| M | 3298 hCoV-19/Australia/VIC15704/2020\|EPI_ISL_593416\|202 | ···················· | ···················· | ···················· | ···················· | ···················· | ···················· | |
| M | 3299 hCoV-19/Australia/VIC15705/2020\|EPI_ISL_593417\|202 | ···················· | ···················· | ···················· | ···················· | ···················· | ···················· | |
| M | 3300 hCoV-19/Australia/VIC15708/2020\|EPI_ISL_593418\|202 | ···················· | ···················· | ···················· | ···················· | ···················· | ···················· | |
| M | 3301 hCoV-19/Australia/VIC15713/2020\|EPI_ISL_593419\|202 | ···················· | ···················· | ···················· | ···················· | ···················· | ···················· | |
| M | 3302 hCoV-19/Australia/VIC15715/2020\|EPI_ISL_593420\|202 | ···················· | ···················· | ···················· | ···················· | ···················· | ···················· | |
| M | 3304 hCoV-19/Australia/VIC15718/2020\|EPI_ISL_593422\|202 | ···················· | ···················· | ···················· | ···················· | ···················· | ···················· | |
| M | 3305 hCoV-19/Australia/VIC15723/2020\|EPI_ISL_593423\|202 | ···················· | ······A············· | ···················· | ···················· | ···················· | ···················· | |
| M | 3308 hCoV-19/Australia/VIC15727/2020\|EPI_ISL_593426\|202 | ···················· | ···················· | ···················· | ···················· | ···················· | ···················· | |
| M | 3310 hCoV-19/Australia/VIC15730/2020\|EPI_ISL_593428\|202 | ···················· | ···················· | ···················· | ···················· | ···················· | ···················· | |
| M | 3311 hCoV-19/Australia/VIC15731/2020\|EPI_ISL_593429\|202 | ···················· | ···················· | ···················· | ···················· | ···················· | ···················· | |
| M | 3312 hCoV-19/USA/WA-ISB1/2020\|EPI_ISL_593596\|2020-07 | ···················· | ···················· | ···················· | ···················· | ···················· | ···················· | |
| M | 3313 hCoV-19/Australia/QLD1078/2020\|EPI_ISL_593597\|2020 | ···················· | ···················· | ···················· | ···················· | ···················· | ···················· | |
| M | 3314 hCoV-19/Australia/QLD1079/2020\|EPI_ISL_593598\|2020 | ···················· | ···················· | ···················· | ···················· | ···················· | ···················· | |
| M | 3315 hCoV-19/Australia/QLD1080/2020\|EPI_ISL_593599\|2020 | ···················· | ···················· | ···················· | ···················· | ···················· | ···················· | |
| M | 3316 hCoV-19/Scotland/CVR4959/2020\|EPI_ISL_594723\|2020- | ···················· | ···················· | ···················· | ···················· | ···················· | ···················· | |
| M | 3317 hCoV-19/Scotland/CVR4960/2020\|EPI_ISL_594724\|2020- | ···················· | ···················· | ···················· | ···················· | ···················· | NN·················· | |
| M | 3318 hCoV-19/Scotland/CVR4965/2020\|EPI_ISL_594729\|2020- | ···················· | ···················· | ···················· | ···················· | ···················· | ···················· | |
| M | 3319 hCoV-19/Scotland/CVR4977/2020\|EPI_ISL_594740\|2020- | ···················· | ···················· | ···················· | ···················· | ···················· | ···················· | |
| M | 3320 hCoV-19/Scotland/CVR4979/2020\|EPI_ISL_594741\|2020- | ···················· | ···················· | ···················· | ···················· | ··············NN···· | NN·N················ | 1 |
| M | 3321 hCoV-19/Scotland/CVR5003/2020\|EPI_ISL_594742\|2020- | ···················· | ···················· | ···················· | ···················· | ···················· | ···················· | |
| M | 3322 hCoV-19/Scotland/CVR5005/2020\|EPI_ISL_594744\|2020- | ···················· | ···················· | ···················· | ···················· | ···················· | ···················· | |
| M | 3323 hCoV-19/Scotland/CVR5006/2020\|EPI_ISL_594745\|2020- | ···················· | ···················· | ···················· | ···················· | ···················· | ···················· | |
| M | 3324 hCoV-19/Scotland/CVR5010/2020\|EPI_ISL_594749\|2020- | ···················· | ···················· | ···················· | ···················· | ···················· | ···················· | |
| M | 3325 hCoV-19/Scotland/CVR5023/2020\|EPI_ISL_594757\|2020- | ···················· | ···················· | ···················· | ···················· | ···················· | ···················· | |
| M | 3326 hCoV-19/Scotland/CVR5024/2020\|EPI_ISL_594758\|2020- | ···················· | ···················· | ···················· | ···················· | ···················· | ···················· | |
| M | 3327 hCoV-19/Scotland/CVR5025/2020\|EPI_ISL_594759\|2020- | ···················· | ···················· | ···················· | ···················· | ·NN················· | NN·····N············ | 1 |
| M | 3328 hCoV-19/Scotland/CVR5026/2020\|EPI_ISL_594760\|2020- | ···················· | ···················· | ···················· | ···················· | ···················· | ···················· | |
| M | 3329 hCoV-19/England/SHEF-C187C/2020\|EPI_ISL_585010\|202 | ···················· | ···················· | ···················· | ···················· | ···················· | ···················· | |
| M | 3330 hCoV-19/England/SHEF-CA3CD/2020\|EPI_ISL_585039\|202 | ···················· | ···················· | ···················· | ···················· | ···················· | ···················· | |
| M | 3331 hCoV-19/England/SHEF-CAC86/2020\|EPI_ISL_585042\|202 | ···················· | ···················· | ···················· | ···················· | ···················· | ···················· | |
| M | 3334 hCoV-19/England/SHEF-CBF13/2020\|EPI_ISL_585046\|202 | ···················· | ···················· | ···················· | ···················· | ···················· | ···················· | |
| M | 3336 hCoV-19/England/SHEF-CC659/2020\|EPI_ISL_585049\|202 | ···················· | ···················· | ···················· | ···················· | ···················· | ···················· | |
| M | 3337 hCoV-19/England/SHEF-CC808/2020\|EPI_ISL_585050\|202 | ···················· | ···················· | ···················· | ···················· | ···················· | ···················· | |
| M | 3338 hCoV-19/England/SHEF-CC914/2020\|EPI_ISL_585051\|202 | ···················· | ·A·················· | ···················· | ···················· | ···················· | ···A················ | |
| M | 3340 hCoV-19/England/SHEF-CCE42/2020\|EPI_ISL_585054\|202 | ···················· | ···················· | ···················· | ···················· | ···················· | ···················· | |
| M | 3342 hCoV-19/England/SHEF-CD281/2020\|EPI_ISL_585058\|202 | ···················· | ···················· | ···················· | ···················· | ···················· | ···················· | |
| M | 3343 hCoV-19/Canada/ON-CVD00726/2020\|EPI_ISL_586271\|202 | ···················· | ···················· | ···················· | ···················· | ···················· | ···················· | |
| M | 3344 hCoV-19/Canada/ON-CVD01006/2020\|EPI_ISL_586274\|202 | ···················· | ···················· | ···················· | ···················· | ···················· | ···················· | |
| M | 3345 hCoV-19/Canada/ON-CVD01345/2020\|EPI_ISL_586276\|202 | ···················· | ···················· | ···················· | ···················· | ···················· | ···················· | |
| M | 3346 hCoV-19/Canada/ON-S1013/2020\|EPI_ISL_586281\|2020-0 | ···················· | ···················· | ···················· | ···················· | ···················· | ···················· | |
| M | 3347 hCoV-19/Canada/ON-S120/2020\|EPI_ISL_586293\|2020-03 | ···················· | ···················· | ···················· | ···················· | ···················· | ···················· | |
| M | 3348 hCoV-19/Canada/ON-S1219/2020\|EPI_ISL_586295\|2020-0 | ···················· | ···················· | ···················· | ···················· | ···················· | ···················· | |
| M | 3349 hCoV-19/Canada/ON-S1220/2020\|EPI_ISL_586297\|2020-0 | ···················· | ···················· | ···················· | ···················· | ···················· | ···················· | |
| M | 3350 hCoV-19/Canada/ON-S1222/2020\|EPI_ISL_586299\|2020-0 | ···················· | ···················· | ···················· | ···················· | ···················· | ···················· | |
| M | 3351 hCoV-19/Canada/ON-S1231/2020\|EPI_ISL_586306\|2020-0 | ···················· | ···················· | ···················· | ···················· | ···················· | ···················· | |
| M | 3352 hCoV-19/Canada/ON-S1238/2020\|EPI_ISL_586309\|2020-0 | ···················· | ···················· | ···················· | ···················· | ···················· | ···················· | |
| M | 3353 hCoV-19/Canada/ON-S1242/2020\|EPI_ISL_586310\|2020-0 | ···················· | ···················· | ···················· | ···················· | ···················· | ···················· | |
| M | 3354 hCoV-19/Canada/ON-S1243/2020\|EPI_ISL_586311\|2020-0 | ···················· | ···················· | ···················· | ···················· | ···················· | ···················· | |
| M | 3355 hCoV-19/Canada/ON-S1257/2020\|EPI_ISL_586316\|2020-0 | ···················· | ···················· | ···················· | ···················· | ···················· | ···················· | |
| M | 3356 hCoV-19/Canada/ON-S1264/2020\|EPI_ISL_586317\|2020-0 | ···················· | ··········A········· | ···················· | ···················· | ···················· | ···················· | |
| M | 3357 hCoV-19/Canada/ON-S1266/2020\|EPI_ISL_586319\|2020-0 | ···················· | ···················· | ···················· | ···················· | ···················· | ···················· | |

| | | | | | | | | |
|---|---|---|---|---|---|---|---|---|
| M | 3358 | hCoV-19/Canada/ON-S1268/2020\|EPI_ISL_586320\|2020-0 | ···················· | ···················· | ···················· | ···················· | ···················· | ···················· |
| M | 3359 | hCoV-19/Canada/ON-S1272/2020\|EPI_ISL_586324\|2020-0 | ···················· | ···················· | ···················· | ···················· | ···················· | ···················· |
| M | 3360 | hCoV-19/Canada/ON-S1274/2020\|EPI_ISL_586325\|2020-0 | ···················· | ···················· | ···················· | ···················· | ···················· | ···················· |
| M | 3361 | hCoV-19/Canada/ON-S1275/2020\|EPI_ISL_586326\|2020-0 | ···················· | ···················· | ···················· | ···················· | ···················· | ···················· |
| M | 3362 | hCoV-19/Canada/ON-S1277/2020\|EPI_ISL_586328\|2020-0 | ···················· | ···················· | ···················· | ···················· | ···················· | ···················· |
| M | 3363 | hCoV-19/Canada/ON-S1278/2020\|EPI_ISL_586329\|2020-0 | ···················· | ···················· | ···················· | ···················· | ···················· | ···················· |
| M | 3364 | hCoV-19/Canada/ON-S1280/2020\|EPI_ISL_586330\|2020-0 | ···················· | ···················· | ···················· | ···················· | ···················· | ···················· |
| M | 3365 | hCoV-19/Canada/ON-S1283/2020\|EPI_ISL_586332\|2020-0 | ···················· | ···················· | ···················· | ···················· | ···················· | ···················· |
| M | 3366 | hCoV-19/Canada/ON-S1285/2020\|EPI_ISL_586333\|2020-0 | ···················· | ···················· | ···················· | ···················· | ···················· | ···················· |
| M | 3367 | hCoV-19/Canada/ON-S1286/2020\|EPI_ISL_586334\|2020-0 | ···················· | ···················· | ···················· | ···················· | ···················· | ···················· |
| M | 3368 | hCoV-19/Canada/ON-S1288/2020\|EPI_ISL_586335\|2020-0 | ···················· | ···················· | ···················· | ···················· | ···················· | ···················· |
| M | 3369 | hCoV-19/Canada/ON-S129/2020\|EPI_ISL_586336\|2020-03 | ···················· | ···················· | ···················· | ···················· | ···················· | ···················· |
| M | 3370 | hCoV-19/Canada/ON-S1292/2020\|EPI_ISL_586338\|2020-0 | ···················· | ···················· | ···················· | ···················· | ···················· | ···················· |
| M | 3371 | hCoV-19/Canada/ON-S1297/2020\|EPI_ISL_586341\|2020-0 | ···················· | ···················· | ···················· | ···················· | ···················· | ···················· |
| M | 3372 | hCoV-19/Canada/ON-S189/2020\|EPI_ISL_586376\|2020-03 | ···················· | ···················· | ···················· | ···················· | ···················· | ···················· |
| M | 3373 | hCoV-19/Canada/ON-S197/2020\|EPI_ISL_586378\|2020-03 | ···················· | ···················· | ···················· | ···················· | ···················· | ···················· |
| M | 3374 | hCoV-19/Canada/ON-S215/2020\|EPI_ISL_586380\|2020-03 | ···················· | ···················· | ···················· | ···················· | ···················· | ···················· |
| M | 3375 | hCoV-19/Canada/ON-S234/2020\|EPI_ISL_586387\|2020-03 | ···················· | ···················· | ···················· | ···················· | ···················· | ···················· |
| M | 3376 | hCoV-19/England/QEUH-A1FCE2/2020\|EPI_ISL_587633\|20 | ···················· | ···················· | ···················· | ···················· | ···················· | ···················· |
| M | 3377 | hCoV-19/England/QEUH-A2101E/2020\|EPI_ISL_587634\|20 | ···················· | ···················· | ···················· | ···················· | ···················· | ···················· |
| M | 3378 | hCoV-19/England/QEUH-A21272/2020\|EPI_ISL_587635\|20 | ···················· | ···················· | ···················· | ···················· | ···················· | ···················· |
| M | 3379 | hCoV-19/England/QEUH-A204E6/2020\|EPI_ISL_587636\|20 | ···················· | ···················· | ···················· | ···················· | ···················· | ···················· |
| M | 3380 | hCoV-19/England/QEUH-A21DF9/2020\|EPI_ISL_587637\|20 | ···················· | ···················· | ···················· | ···················· | ······T············· | ···················· |
| M | 3381 | hCoV-19/England/QEUH-A2052F/2020\|EPI_ISL_587638\|20 | ···················· | ···················· | ···················· | ···················· | ···················· | ···················· |
| M | 3382 | hCoV-19/England/QEUH-A2245D/2020\|EPI_ISL_587640\|20 | ···················· | ···················· | ···················· | ···················· | ···················· | ···················· |
| M | 3383 | hCoV-19/England/QEUH-A20273/2020\|EPI_ISL_587641\|20 | ···················· | ···················· | ···················· | ···················· | ···················· | ···················· |
| M | 3384 | hCoV-19/England/QEUH-A21E41/2020\|EPI_ISL_587642\|20 | ···················· | ···················· | ···················· | ···················· | ···················· | ···················· |
| M | 3385 | hCoV-19/England/QEUH-A215B5/2020\|EPI_ISL_587643\|20 | ···················· | ···················· | ···················· | ···················· | ···················· | ···················· |
| M | 3386 | hCoV-19/England/QEUH-A20291/2020\|EPI_ISL_587644\|20 | ···················· | ···················· | ···················· | ···················· | ···················· | ···················· |
| M | 3387 | hCoV-19/England/QEUH-A2047D/2020\|EPI_ISL_587645\|20 | ···················· | ···················· | ···················· | ···················· | ···················· | ···················· |
| M | 3388 | hCoV-19/England/QEUH-A21236/2020\|EPI_ISL_587646\|20 | ···················· | ·A·················· | ···················· | ···················· | ···················· | ···················· |
| M | 3389 | hCoV-19/England/QEUH-A2099C/2020\|EPI_ISL_587647\|20 | ···················· | ···················· | ···················· | ···················· | ···················· | ···················· |
| M | 3390 | hCoV-19/England/QEUH-A21E32/2020\|EPI_ISL_587648\|20 | ···················· | ···················· | ···················· | ···················· | ···················· | ···················· |
| M | 3391 | hCoV-19/England/QEUH-A202BF/2020\|EPI_ISL_587649\|20 | ···················· | ···················· | ···················· | ···················· | ···················· | ···················· |
| M | 3392 | hCoV-19/England/QEUH-A21281/2020\|EPI_ISL_587650\|20 | ···················· | ···················· | ···················· | ···················· | ···················· | ···················· |
| M | 3393 | hCoV-19/England/QEUH-A2055C/2020\|EPI_ISL_587651\|20 | ···················· | ···················· | ···················· | ···················· | ···················· | ···················· |
| M | 3394 | hCoV-19/England/QEUH-A202DD/2020\|EPI_ISL_587652\|20 | ···················· | ···················· | ···················· | ···················· | ···················· | ···················· |
| M | 3395 | hCoV-19/England/QEUH-A209D8/2020\|EPI_ISL_587653\|20 | ···················· | ···················· | ···················· | ···················· | ···················· | ···················· |
| M | 3396 | hCoV-19/England/QEUH-A20282/2020\|EPI_ISL_587655\|20 | ···················· | ···················· | ···················· | ···················· | ···················· | ···················· |
| M | 3397 | hCoV-19/England/QEUH-A20747/2020\|EPI_ISL_587656\|20 | ···················· | ···················· | ···················· | ···················· | ···················· | ···················· |
| M | 3398 | hCoV-19/England/QEUH-A21EAB/2020\|EPI_ISL_587657\|20 | ···················· | ···················· | ···················· | ···················· | ···················· | ···················· |
| M | 3399 | hCoV-19/England/QEUH-A2160D/2020\|EPI_ISL_587658\|20 | ···················· | ···················· | ···················· | ···················· | ···················· | ···················· |
| M | 3400 | hCoV-19/England/QEUH-A20729/2020\|EPI_ISL_587659\|20 | ···················· | ···················· | ···················· | ···················· | ···················· | ···················· |
| M | 3401 | hCoV-19/England/QEUH-A21E50/2020\|EPI_ISL_587660\|20 | ···················· | ···················· | ···················· | ···················· | ···················· | ···················· |
| M | 3402 | hCoV-19/England/QEUH-A20264/2020\|EPI_ISL_587661\|20 | ···················· | ···················· | ···················· | ···················· | ···················· | ···················· |
| M | 3403 | hCoV-19/England/QEUH-A2098D/2020\|EPI_ISL_587662\|20 | ···················· | ···················· | ···················· | ···················· | ···················· | ···················· |
| M | 3404 | hCoV-19/England/QEUH-A1FC88/2020\|EPI_ISL_587663\|20 | ···················· | ···················· | ···················· | ···················· | ···················· | ···················· |
| M | 3405 | hCoV-19/England/QEUH-A21E9C/2020\|EPI_ISL_587664\|20 | ···················· | ···················· | ···················· | ···················· | ···················· | ···················· |
| M | 3406 | hCoV-19/England/QEUH-A2161C/2020\|EPI_ISL_587665\|20 | ···················· | ···················· | ···················· | ···················· | ····T··············· | ···················· |
| M | 3407 | hCoV-19/England/QEUH-A212BE/2020\|EPI_ISL_587667\|20 | ···················· | ···················· | ···················· | ···················· | ···················· | ···················· |
| M | 3408 | hCoV-19/England/QEUH-A2038F/2020\|EPI_ISL_587668\|20 | ···················· | ···················· | ···················· | ···················· | ···················· | ···················· |
| M | 3409 | hCoV-19/England/QEUH-A202EC/2020\|EPI_ISL_587669\|20 | ···················· | ···················· | ···················· | ···················· | ···················· | ···················· |
| M | 3410 | hCoV-19/England/QEUH-A226DF/2020\|EPI_ISL_587670\|20 | ···················· | ···················· | ···················· | ···················· | ···················· | ···················· |
| M | 3411 | hCoV-19/England/QEUH-A204B9/2020\|EPI_ISL_587671\|20 | ···················· | ···················· | ···················· | ···················· | ···················· | ···················· |
| M | 3412 | hCoV-19/England/QEUH-A2048C/2020\|EPI_ISL_587672\|20 | ···················· | ···················· | ···················· | ···················· | ···················· | ···················· |
| M | 3413 | hCoV-19/England/QEUH-A21E05/2020\|EPI_ISL_587673\|20 | ···················· | ···················· | ···················· | ···················· | ···················· | ···················· |
| M | 3414 | hCoV-19/England/QEUH-A22420/2020\|EPI_ISL_587674\|20 | ···················· | ···················· | ···················· | ···················· | ···················· | ···················· |
| M | 3415 | hCoV-19/England/QEUH-A21E8D/2020\|EPI_ISL_587676\|20 | ···················· | ···················· | ···················· | ···················· | ····T··············· | ···················· |
| M | 3416 | hCoV-19/England/QEUH-A1F9EB/2020\|EPI_ISL_587699\|20 | ···················· | ···················· | ···················· | ···················· | ···················· | ···················· |
| M | 3417 | hCoV-19/England/QEUH-9F805D/2020\|EPI_ISL_588965\|20 | ···················· | ···················· | ···················· | ···················· | ···················· | ···················· |
| M | 3418 | hCoV-19/Wales/QEUH-9F93BD/2020\|EPI_ISL_588966\|2020 | ···················· | ···················· | ···················· | ···················· | ···················· | ···················· |
| M | 3419 | hCoV-19/England/QEUH-9F81A5/2020\|EPI_ISL_588967\|20 | ···················· | ···················· | ···················· | ···················· | ···················· | ···················· |
| M | 3420 | hCoV-19/England/QEUH-9F5700/2020\|EPI_ISL_588968\|20 | ···················· | ···················· | ···················· | ···················· | ···················· | ···················· |
| M | 3421 | hCoV-19/England/QEUH-9F607D/2020\|EPI_ISL_588969\|20 | ···················· | ···················· | ···················· | ···················· | ···················· | ···················· |
| M | 3422 | hCoV-19/England/QEUH-9F861F/2020\|EPI_ISL_588970\|20 | ···················· | ····A··············· | ···················· | ···················· | ···················· | ···················· |
| M | 3423 | hCoV-19/England/QEUH-9F8266/2020\|EPI_ISL_588971\|20 | ···················· | ···················· | ···················· | ···················· | ···················· | ···················· |
| M | 3424 | hCoV-19/England/QEUH-9F7FAC/2020\|EPI_ISL_588972\|20 | ···················· | ···················· | ···················· | ···················· | ···················· | ···················· |
| M | 3425 | hCoV-19/England/QEUH-9F62B3/2020\|EPI_ISL_588973\|20 | ···················· | ···················· | ···················· | ···················· | ···················· | ···················· |
| M | 3426 | hCoV-19/England/QEUH-9F5AF8/2020\|EPI_ISL_588974\|20 | ···················· | ···················· | ···················· | ···················· | ······T············· | ···················· |
| M | 3427 | hCoV-19/England/QEUH-9F6101/2020\|EPI_ISL_588975\|20 | ···················· | ···················· | ···················· | ···················· | ···················· | ···················· |
| M | 3428 | hCoV-19/England/QEUH-9F7CE1/2020\|EPI_ISL_588976\|20 | ···················· | ···················· | ···················· | ···················· | ······T············· | ···················· |
| M | 3429 | hCoV-19/England/QEUH-9F82FD/2020\|EPI_ISL_588977\|20 | ···················· | ···················· | ···················· | ···················· | ···················· | ···················· |
| M | 3430 | hCoV-19/England/QEUH-9F907A/2020\|EPI_ISL_588978\|20 | ···················· | ···················· | ···················· | ···················· | ····T··············· | ···················· |
| M | 3431 | hCoV-19/England/QEUH-9F568B/2020\|EPI_ISL_588979\|20 | ···················· | ·A·················· | ···················· | ···················· | ···················· | ···A················ |
| M | 3432 | hCoV-19/England/QEUH-9F57C4/2020\|EPI_ISL_588980\|20 | ···················· | ·A·················· | ···················· | ···················· | ···················· | ···················· |
| M | 3433 | hCoV-19/England/QEUH-9F8E17/2020\|EPI_ISL_588981\|20 | ···················· | ···················· | ···················· | ···················· | ···················· | ···················· |
| M | 3434 | hCoV-19/England/QEUH-9F8363/2020\|EPI_ISL_588982\|20 | ···················· | ···················· | ···················· | ···················· | ···················· | ···················· |
| M | 3435 | hCoV-19/England/QEUH-9F83AF/2020\|EPI_ISL_588983\|20 | ···················· | ···················· | ···················· | ···················· | ···················· | ···················· |
| M | 3436 | hCoV-19/England/QEUH-9F8A22/2020\|EPI_ISL_588984\|20 | ···················· | ·A·················· | ···················· | ···················· | ···················· | ···A················ |
| M | 3437 | hCoV-19/England/QEUH-9F5F26/2020\|EPI_ISL_588985\|20 | ···················· | ···················· | ···················· | ···················· | ······T············· | ···················· |

| | | | | | | | | |
|---|---|---|---|---|---|---|---|---|
| M | 3438 | hCoV-19/Wales/QEUH-9F6374/2020\|EPI_ISL_588986\|2020 | ···················· | ···················· | ···················· | ···················· | ···················· | ···················· |
| M | 3439 | hCoV-19/England/QEUH-9F87A3/2020\|EPI_ISL_588987\|20 | ···················· | ···················· | ···················· | ···················· | ···················· | ···················· |
| M | 3440 | hCoV-19/England/QEUH-9F93AE/2020\|EPI_ISL_588989\|20 | ···················· | ···················· | ···················· | ···················· | ···················· | ···················· |
| M | 3441 | hCoV-19/England/QEUH-9F8961/2020\|EPI_ISL_588990\|20 | ···················· | ···················· | ···················· | ···················· | ···················· | ···················· |
| M | 3442 | hCoV-19/England/QEUH-9F822A/2020\|EPI_ISL_588991\|20 | ···················· | ···················· | ···················· | ···················· | ···················· | ···················· |
| M | 3443 | hCoV-19/England/QEUH-9F93DB/2020\|EPI_ISL_588993\|20 | ···················· | ·A·················· | ···················· | ···················· | ···················· | ··A················· |
| M | 3444 | hCoV-19/England/QEUH-9F56E5/2020\|EPI_ISL_588994\|20 | ···················· | ···················· | ···················· | ···················· | ···················· | ···················· |
| M | 3445 | hCoV-19/England/QEUH-9F5A16/2020\|EPI_ISL_588996\|20 | ···················· | ···················· | ···················· | ···················· | ···················· | ···················· |
| M | 3446 | hCoV-19/England/QEUH-9F8293/2020\|EPI_ISL_588997\|20 | ···················· | ···················· | ···················· | ···················· | ···················· | ···················· |
| M | 3447 | hCoV-19/England/QEUH-9F87EF/2020\|EPI_ISL_588998\|20 | ···················· | ···················· | ···················· | ···················· | ···················· | ···················· |
| M | 3448 | hCoV-19/England/QEUH-9F8873/2020\|EPI_ISL_588999\|20 | ···················· | ···················· | ···················· | ···················· | ···················· | ···················· |
| M | 3449 | hCoV-19/Australia/VIC12881/2020\|EPI_ISL_592100\|202 | ···················· | ···················· | ···················· | ···················· | ···················· | ···················· |
| M | 3450 | hCoV-19/Australia/VIC12893/2020\|EPI_ISL_592101\|202 | ···················· | ···················· | ···················· | ···················· | ···················· | ···················· |
| M | 3451 | hCoV-19/Australia/VIC12897/2020\|EPI_ISL_592102\|202 | ···················· | ···················· | ···················· | ···················· | ···················· | ···················· |
| M | 3452 | hCoV-19/Australia/VIC12901/2020\|EPI_ISL_592103\|202 | ···················· | ···················· | ···················· | ···················· | ···················· | ···················· |
| M | 3453 | hCoV-19/Australia/VIC12902/2020\|EPI_ISL_592104\|202 | ···················· | ···················· | ···················· | ···················· | ···················· | ···················· |
| M | 3454 | hCoV-19/Australia/VIC12903/2020\|EPI_ISL_592105\|202 | ···················· | ···················· | ···················· | ···················· | ···················· | ···················· |
| M | 3455 | hCoV-19/Australia/VIC12914/2020\|EPI_ISL_592106\|202 | ···················· | ···················· | ···················· | ···················· | ···················· | ···················· |
| M | 3456 | hCoV-19/Australia/VIC12915/2020\|EPI_ISL_592107\|202 | ···················· | ···················· | ···················· | ···················· | ···················· | ···················· |
| M | 3457 | hCoV-19/Australia/VIC12919/2020\|EPI_ISL_592108\|202 | ···················· | ···················· | ···················· | ···················· | ···················· | ···················· |
| M | 3458 | hCoV-19/Australia/VIC12920/2020\|EPI_ISL_592109\|202 | ···················· | ···················· | ···················· | ···················· | ···················· | ···················· |
| M | 3459 | hCoV-19/Australia/VIC12924/2020\|EPI_ISL_592110\|202 | ···················· | ···················· | ···················· | ···················· | ···················· | ···················· |
| M | 3460 | hCoV-19/Australia/VIC12928/2020\|EPI_ISL_592111\|202 | ···················· | ···················· | ···················· | ···················· | ···················· | ···················· |
| M | 3461 | hCoV-19/Australia/VIC12929/2020\|EPI_ISL_592112\|202 | ···················· | ···················· | ···················· | ···················· | ···················· | ···················· |
| M | 3462 | hCoV-19/Australia/VIC12931/2020\|EPI_ISL_592113\|202 | ···················· | ···················· | ···················· | ···················· | ···················· | ···················· |
| M | 3463 | hCoV-19/Australia/VIC12933/2020\|EPI_ISL_592114\|202 | ···················· | ···················· | ···················· | ···················· | ···················· | ···················· |
| M | 3464 | hCoV-19/Australia/VIC12939/2020\|EPI_ISL_592115\|202 | ···················· | ···················· | ···················· | ···················· | ···················· | ···················· |
| M | 3465 | hCoV-19/Australia/VIC12940/2020\|EPI_ISL_592116\|202 | ···················· | ···················· | ···················· | ···················· | ···················· | ···················· |
| M | 3466 | hCoV-19/Australia/VIC12941/2020\|EPI_ISL_592117\|202 | ···················· | ···················· | ···················· | ···················· | ···················· | ···················· |
| M | 3467 | hCoV-19/Australia/VIC12943/2020\|EPI_ISL_592118\|202 | ···················· | ···················· | ···················· | ···················· | ···················· | ···················· |
| M | 3468 | hCoV-19/Australia/VIC12946/2020\|EPI_ISL_592119\|202 | ···················· | ···················· | ···················· | ···················· | ···················· | ···················· |
| M | 3469 | hCoV-19/Australia/VIC12950/2020\|EPI_ISL_592120\|202 | ···················· | ···················· | ···················· | ···················· | ···················· | ···················· |
| M | 3470 | hCoV-19/Australia/VIC12952/2020\|EPI_ISL_592121\|202 | ···················· | ···················· | ···················· | ···················· | ···················· | ···················· |
| M | 3471 | hCoV-19/Australia/VIC12956/2020\|EPI_ISL_592122\|202 | ···················· | ···················· | ···················· | ···················· | ···················· | ···················· |
| M | 3472 | hCoV-19/Australia/VIC12959/2020\|EPI_ISL_592123\|202 | ···················· | ···················· | ···················· | ···················· | ···················· | ···················· |
| M | 3473 | hCoV-19/Australia/VIC12961/2020\|EPI_ISL_592124\|202 | ···················· | ···················· | ···················· | ···················· | ···················· | ···················· |
| M | 3474 | hCoV-19/Australia/VIC12962/2020\|EPI_ISL_592125\|202 | ···················· | ···················· | ···················· | ···················· | ···················· | ···················· |
| M | 3475 | hCoV-19/Australia/VIC12964/2020\|EPI_ISL_592126\|202 | ···················· | ···················· | ···················· | ···················· | ···················· | ···················· |
| M | 3476 | hCoV-19/Australia/VIC12965/2020\|EPI_ISL_592127\|202 | ···················· | ···················· | ···················· | ···················· | ···················· | ···················· |
| M | 3477 | hCoV-19/Australia/VIC12968/2020\|EPI_ISL_592128\|202 | ···················· | ···················· | ···················· | ···················· | ···················· | ···················· |
| M | 3478 | hCoV-19/Australia/VIC12970/2020\|EPI_ISL_592129\|202 | ···················· | ···················· | ···················· | ···················· | ···················· | ···················· |
| M | 3479 | hCoV-19/Australia/VIC12971/2020\|EPI_ISL_592130\|202 | ···················· | ···················· | ···················· | ···················· | ···················· | ···················· |
| M | 3480 | hCoV-19/Australia/VIC12976/2020\|EPI_ISL_592131\|202 | ···················· | ···················· | ···················· | ···················· | ···················· | ···················· |
| M | 3481 | hCoV-19/Australia/VIC12979/2020\|EPI_ISL_592132\|202 | ···················· | ···················· | ···················· | ···················· | ···················· | ···················· |
| M | 3482 | hCoV-19/Australia/VIC12980/2020\|EPI_ISL_592133\|202 | ···················· | ···················· | ···················· | ···················· | ···················· | ···················· |
| M | 3483 | hCoV-19/Australia/VIC12981/2020\|EPI_ISL_592134\|202 | ···················· | ···················· | ···················· | ···················· | ···················· | ···················· |
| M | 3484 | hCoV-19/Australia/VIC12982/2020\|EPI_ISL_592135\|202 | ···················· | ···················· | ···················· | ···················· | ···················· | ···················· |
| M | 3485 | hCoV-19/Australia/VIC12986/2020\|EPI_ISL_592136\|202 | ···················· | ···················· | ··K················· | ···················· | ···················· | ···················· |
| M | 3486 | hCoV-19/Australia/VIC12987/2020\|EPI_ISL_592137\|202 | ···················· | ···················· | ···················· | ···················· | ···················· | ···················· |
| M | 3488 | hCoV-19/Australia/VIC12989/2020\|EPI_ISL_592139\|202 | ···················· | ···················· | ···················· | ···················· | ···················· | ···················· |
| M | 3489 | hCoV-19/Australia/VIC12993/2020\|EPI_ISL_592140\|202 | ···················· | ···················· | ··K················· | ···················· | ···················· | ····M··············· |
| M | 3490 | hCoV-19/Australia/VIC12994/2020\|EPI_ISL_592141\|202 | ···················· | ···················· | ··K················· | ···················· | ···················· | ···················· |
| M | 3491 | hCoV-19/Australia/VIC12995/2020\|EPI_ISL_592142\|202 | ···················· | ···················· | ···················· | ···················· | ···················· | ···················· |
| M | 3492 | hCoV-19/Australia/VIC15659/2020\|EPI_ISL_593400\|202 | ···················· | ···················· | ···················· | ···················· | ···················· | ···················· |
| M | 3493 | hCoV-19/Australia/VIC15660/2020\|EPI_ISL_593401\|202 | ···················· | ···················· | ···················· | ···················· | ···················· | ···················· |
| M | 3494 | hCoV-19/Australia/VIC15662/2020\|EPI_ISL_593402\|202 | ···················· | ···················· | ···················· | ···················· | ···················· | ···················· |
| M | 3495 | hCoV-19/Australia/VIC15663/2020\|EPI_ISL_593403\|202 | ···················· | ···················· | ···················· | ···················· | ···················· | ···················· |
| M | 3496 | hCoV-19/Australia/VIC15664/2020\|EPI_ISL_593404\|202 | ···················· | ···················· | ···················· | ···················· | ···················· | ···················· |
| M | 3497 | hCoV-19/Australia/VIC15666/2020\|EPI_ISL_593405\|202 | ···················· | ···················· | ···················· | ···················· | ···················· | ···················· |
| M | 3499 | hCoV-19/Australia/VIC15671/2020\|EPI_ISL_593407\|202 | ···················· | ···················· | ···················· | ···················· | ···················· | ···················· |
| M | 3500 | hCoV-19/Australia/VIC15672/2020\|EPI_ISL_593408\|202 | ···················· | ···················· | ···················· | ···················· | NNN················· | ···················· | 1
| M | 3501 | hCoV-19/Australia/VIC15674/2020\|EPI_ISL_593409\|202 | ···················· | ···················· | ···················· | ···················· | ···················· | ···················· |
| M | 3502 | hCoV-19/Australia/VIC15732/2020\|EPI_ISL_593430\|202 | ···················· | ···················· | ···················· | ···················· | ···················· | ···················· |
| M | 3507 | hCoV-19/Australia/VIC15741/2020\|EPI_ISL_593435\|202 | ···················· | ···················· | ···················· | ···················· | ···················· | ···················· |
| M | 3508 | hCoV-19/Australia/VIC15743/2020\|EPI_ISL_593436\|202 | ···················· | ···················· | ···················· | ···················· | ···················· | ···················· |
| M | 3510 | hCoV-19/Australia/VIC15746/2020\|EPI_ISL_593438\|202 | ···················· | ···················· | ···················· | ···················· | ···················· | ···················· |
| M | 3511 | hCoV-19/Australia/VIC15750/2020\|EPI_ISL_593439\|202 | ···················· | ···················· | ···················· | ···················· | ···················· | ···················· |
| M | 3512 | hCoV-19/Australia/VIC15752/2020\|EPI_ISL_593440\|202 | ···················· | ···················· | ···················· | ···················· | ···················· | ···················· |
| M | 3515 | hCoV-19/Australia/VIC15756/2020\|EPI_ISL_593443\|202 | ···················· | ···················· | ···················· | ···················· | ···················· | ···················· |
| M | 3516 | hCoV-19/Australia/VIC15757/2020\|EPI_ISL_593444\|202 | ···················· | ···················· | ···················· | ···················· | ···················· | ···················· |
| M | 3517 | hCoV-19/Australia/VIC15758/2020\|EPI_ISL_593445\|202 | ···················· | ···················· | ···················· | ···················· | ···················· | ···················· |
| M | 3518 | hCoV-19/Australia/VIC15760/2020\|EPI_ISL_593446\|202 | ···················· | ···················· | ···················· | ···················· | ···················· | ···················· |
| M | 3519 | hCoV-19/Australia/VIC15763/2020\|EPI_ISL_593447\|202 | ···················· | ···················· | ···················· | ···················· | ···················· | ···················· |
| M | 3520 | hCoV-19/Australia/VIC15765/2020\|EPI_ISL_593448\|202 | ···················· | ···················· | ···················· | ···················· | ···················· | ···················· |
| M | 3522 | hCoV-19/Australia/VIC15772/2020\|EPI_ISL_593450\|202 | ···················· | ···················· | ···················· | ···················· | ···················· | ···················· |
| M | 3523 | hCoV-19/Australia/VIC15773/2020\|EPI_ISL_593451\|202 | ···················· | ···················· | ···················· | ···················· | ···················· | ···················· |
| M | 3527 | hCoV-19/Australia/VIC15781/2020\|EPI_ISL_593455\|202 | ···················· | ···················· | ···················· | ···················· | ···················· | ···················· |
| M | 3528 | hCoV-19/Australia/VIC15783/2020\|EPI_ISL_593456\|202 | ···················· | ···················· | ···················· | ···················· | ···················· | ···················· |
| M | 3529 | hCoV-19/Australia/VIC15784/2020\|EPI_ISL_593457\|202 | ···················· | ···················· | ···················· | ···················· | ···················· | ···················· |
| M | 3530 | hCoV-19/Australia/VIC15786/2020\|EPI_ISL_593458\|202 | ···················· | ···················· | ···················· | ···················· | ···················· | ···················· |

| | | | | | | | | |
|---|---|---|---|---|---|---|---|---|
| M | 3531 hCoV-19/Australia/VIC15789/2020\|EPI_ISL_593459\|202 | .................... | .................... | .................... | .................... | .................... | .................... | |
| M | 3532 hCoV-19/Australia/VIC15800/2020\|EPI_ISL_593460\|202 | .................... | .................... | .................... | .................... | .................... | .................... | |
| M | 3533 hCoV-19/Australia/VIC15803/2020\|EPI_ISL_593461\|202 | .................... | .................... | .................... | .................... | .................... | .................... | |
| M | 3534 hCoV-19/Australia/VIC15811/2020\|EPI_ISL_593462\|202 | .................... | .................... | .................... | .................... | .................... | .................... | |
| M | 3535 hCoV-19/Australia/VIC15813/2020\|EPI_ISL_593463\|202 | .................... | .................... | .................... | .................... | .................... | .................... | |
| M | 3537 hCoV-19/Australia/VIC15816/2020\|EPI_ISL_593465\|202 | .................... | .................... | .................... | .................... | .................... | .................... | |
| M | 3538 hCoV-19/Australia/VIC15818/2020\|EPI_ISL_593466\|202 | .................... | .................... | .................... | .................... | .................... | .................... | |
| M | 3539 hCoV-19/Australia/VIC15823/2020\|EPI_ISL_593467\|202 | .................... | .................... | .................... | .................... | .................... | .................... | |
| M | 3540 hCoV-19/Australia/VIC15828/2020\|EPI_ISL_593468\|202 | .................... | .................... | .................... | .................... | .................... | .................... | |
| M | 3541 hCoV-19/Australia/VIC15830/2020\|EPI_ISL_593469\|202 | .................... | .................... | .................... | .................... | .................... | .................... | |
| M | 3542 hCoV-19/Australia/VIC15832/2020\|EPI_ISL_593470\|202 | .................... | .................... | .................... | .................... | ...R................ | .................... | |
| M | 3543 hCoV-19/Australia/VIC15835/2020\|EPI_ISL_593471\|202 | .................... | .................... | .................... | .................... | .................... | .................... | |
| M | 3544 hCoV-19/Australia/VIC15836/2020\|EPI_ISL_593472\|202 | .................... | .................... | .................... | .................... | .................... | .................... | |
| M | 3546 hCoV-19/Scotland/CVR4934/2020\|EPI_ISL_594700\|2020- | .................... | .................... | .................... | .................... | .................... | .................... | |
| M | 3547 hCoV-19/Scotland/CVR4935/2020\|EPI_ISL_594701\|2020- | .................... | .................... | .................... | .................... | .................... | .................... | |
| M | 3549 hCoV-19/Scotland/CVR4937/2020\|EPI_ISL_594703\|2020- | .................... | .................... | .................... | .................... | .................... | .................... | |
| M | 3550 hCoV-19/Scotland/CVR4939/2020\|EPI_ISL_594705\|2020- | .................... | .................... | .................... | .................... | .................... | .................... | |
| M | 3551 hCoV-19/Scotland/CVR4942/2020\|EPI_ISL_594706\|2020- | .................... | .................... | .................... | .................... | ..............N..... | .................... | |
| M | 3552 hCoV-19/Scotland/CVR4947/2020\|EPI_ISL_594711\|2020- | .................... | .................... | .................... | .................... | .................... | .................... | |
| M | 3553 hCoV-19/Scotland/CVR4950/2020\|EPI_ISL_594714\|2020- | .................... | .................... | .................... | .................... | .................... | .................... | |
| M | 3554 hCoV-19/Scotland/CVR4951/2020\|EPI_ISL_594715\|2020- | .................... | .................... | .................... | .................... | .................... | .................... | |
| M | 3555 hCoV-19/Scotland/CVR4952/2020\|EPI_ISL_594716\|2020- | .................... | .................... | .................... | .................... | .................... | .................... | |
| M | 3556 hCoV-19/Scotland/CVR4955/2020\|EPI_ISL_594719\|2020- | .................... | .................... | .................... | .................... | .................... | .................... | |
| M | 3557 hCoV-19/Scotland/CVR4964/2020\|EPI_ISL_594728\|2020- | .................... | .................... | .................... | .................... | .................... | .................... | |
| M | 3559 hCoV-19/Scotland/CVR4967/2020\|EPI_ISL_594731\|2020- | .................... | .................... | .................... | .................... | .................... | .................... | |
| M | 3560 hCoV-19/Scotland/CVR4972/2020\|EPI_ISL_594735\|2020- | .................... | .................... | .................... | .................... | .................... | .................... | |
| M | 3562 hCoV-19/Scotland/CVR4975/2020\|EPI_ISL_594738\|2020- | .................... | .................... | .................... | .................... | .................... | .................... | |
| M | 3563 hCoV-19/Scotland/CVR4976/2020\|EPI_ISL_594739\|2020- | .................... | .................... | .................... | .................... | .................... | .................... | |
| M | 3564 hCoV-19/Scotland/CVR5027/2020\|EPI_ISL_594761\|2020- | .................... | .................... | .................... | .................... | .................... | .................... | |
| M | 3565 hCoV-19/Scotland/CVR5033/2020\|EPI_ISL_594767\|2020- | .................... | .................... | .................... | .................... | .................... | .................... | |
| M | 3566 hCoV-19/Scotland/CVR5037/2020\|EPI_ISL_594771\|2020- | .................... | .................... | .................... | .................... | .................... | .................... | |
| M | 3567 hCoV-19/Scotland/CVR5038/2020\|EPI_ISL_594772\|2020- | .................... | .................... | .................... | .................... | .................... | NN.........N........ | 1 |
| M | 3568 hCoV-19/Scotland/CVR5039/2020\|EPI_ISL_594773\|2020- | .................... | .................... | .................... | .................... | .................... | .................... | |
| M | 3569 hCoV-19/Scotland/CVR5040/2020\|EPI_ISL_594774\|2020- | .................... | .................... | .................... | .................... | .................... | .................... | |
| M | 3570 hCoV-19/Scotland/CVR5043/2020\|EPI_ISL_594777\|2020- | .................... | .................... | .................... | .................... | ..............N..... | NN.................. | |
| M | 3571 hCoV-19/Scotland/CVR5044/2020\|EPI_ISL_594778\|2020- | .................... | .................... | .................... | .................... | .................... | .................... | |
| M | 3572 hCoV-19/Scotland/CVR5045/2020\|EPI_ISL_594779\|2020- | .................... | .................... | .................... | .................... | .................... | .................... | |
| M | 3573 hCoV-19/Scotland/CVR5046/2020\|EPI_ISL_594780\|2020- | .................... | .................... | .................... | .................... | .................... | NN.................. | |
| M | 3574 hCoV-19/Scotland/CVR5047/2020\|EPI_ISL_594781\|2020- | .................... | .................... | .................... | .................... | .................... | .................... | |
| M | 3575 hCoV-19/Scotland/CVR5049/2020\|EPI_ISL_594783\|2020- | .................... | .................... | .................... | .................... | .................... | .................... | |
| M | 3576 hCoV-19/Scotland/CVR5050/2020\|EPI_ISL_594784\|2020- | .................... | .................... | .................... | .................... | .................... | .................... | |
| M | 3577 hCoV-19/Scotland/CVR5051/2020\|EPI_ISL_594785\|2020- | .................... | .................... | .................... | .................... | .................... | .................... | |
| M | 3578 hCoV-19/Scotland/CVR5055/2020\|EPI_ISL_594789\|2020- | .................... | .................... | .................... | .................... | .................... | .................... | |
| M | 3579 hCoV-19/Scotland/CVR5056/2020\|EPI_ISL_594790\|2020- | .................... | .................... | .................... | .................... | .................... | .................... | |
| M | 3580 hCoV-19/Scotland/CVR5060/2020\|EPI_ISL_594793\|2020- | .................... | .................... | .................... | .................... | .................... | .................... | |
| M | 3581 hCoV-19/Scotland/CVR5062/2020\|EPI_ISL_594795\|2020- | .................... | .................... | .................... | .................... | .................... | .................... | |
| M | 3582 hCoV-19/Scotland/CVR5063/2020\|EPI_ISL_594796\|2020- | .................... | .................... | .................... | .................... | .................... | .................... | |
| M | 3583 hCoV-19/Scotland/CVR5064/2020\|EPI_ISL_594797\|2020- | .................... | .................... | .................... | .................... | .................... | .................... | |
| M | 3584 hCoV-19/Scotland/CVR5065/2020\|EPI_ISL_594798\|2020- | .................... | .................... | .................... | .................... | .................... | .................... | |
| M | 3585 hCoV-19/England/204180127/2020\|EPI_ISL_583972\|2020 | .................... | .................... | .................... | .................... | .................... | .................... | |
| M | 3586 hCoV-19/England/204180130/2020\|EPI_ISL_583974\|2020 | .................... | .................... | .................... | .................... | .................... | .................... | |
| M | 3587 hCoV-19/England/204180132/2020\|EPI_ISL_583976\|2020 | .................... | .................... | .................... | .................... | .................... | .................... | |
| M | 3588 hCoV-19/England/204180134/2020\|EPI_ISL_583977\|2020 | .................... | .................... | .................... | .................... | .................... | .................... | |
| M | 3589 hCoV-19/England/204180135/2020\|EPI_ISL_583978\|2020 | .................... | .................... | .................... | .................... | .................... | .................... | |
| M | 3590 hCoV-19/England/204180138/2020\|EPI_ISL_583981\|2020 | .................... | .................... | .................... | .................... | .................... | .................... | |
| M | 3591 hCoV-19/England/204180140/2020\|EPI_ISL_583983\|2020 | .................... | .................... | .................... | .................... | .................... | .................... | |
| M | 3592 hCoV-19/England/204180141/2020\|EPI_ISL_583984\|2020 | .................... | .................... | .................... | .................... | .................... | .................... | |
| M | 3593 hCoV-19/England/204180142/2020\|EPI_ISL_583985\|2020 | .................... | .................... | .................... | .................... | .................... | .................... | |
| M | 3594 hCoV-19/England/204180143/2020\|EPI_ISL_583986\|2020 | .................... | .................... | .................... | .................... | .................... | .................... | |
| M | 3595 hCoV-19/England/204180253/2020\|EPI_ISL_583987\|2020 | .................... | .................... | .................... | .................... | .................... | .................... | |
| M | 3596 hCoV-19/England/204180254/2020\|EPI_ISL_583988\|2020 | .................... | .................... | .................... | .................... | .................... | .................... | |
| M | 3597 hCoV-19/England/204180255/2020\|EPI_ISL_583989\|2020 | .................... | .................... | .................... | .................... | .................... | .................... | |
| M | 3598 hCoV-19/England/204180258/2020\|EPI_ISL_583990\|2020 | .................... | .................... | .................... | .................... | .................... | .................... | |
| M | 3599 hCoV-19/England/204180259/2020\|EPI_ISL_583991\|2020 | .................... | .................... | .................... | .................... | .................... | .................... | |
| M | 3600 hCoV-19/England/SHEF-C37D7/2020\|EPI_ISL_585018\|202 | .................... | .................... | .................... | .................... | .................... | .................... | |
| M | 3602 hCoV-19/England/SHEF-C54ED/2020\|EPI_ISL_585020\|202 | .................... | .................... | .................... | .................... | .................... | .................... | |
| M | 3603 hCoV-19/England/SHEF-C55F9/2020\|EPI_ISL_585022\|202 | .................... | .................... | .................... | .................... | .................... | .................... | |
| M | 3604 hCoV-19/England/SHEF-C5720/2020\|EPI_ISL_585023\|202 | .................... | .................... | .................... | .................... | .................... | .................... | |
| M | 3606 hCoV-19/England/SHEF-C70C9/2020\|EPI_ISL_585028\|202 | .................... | .................... | .................... | .................... | .................... | .................... | |
| M | 3607 hCoV-19/England/SHEF-C70E7/2020\|EPI_ISL_585029\|202 | .................... | .................... | .................... | .................... | .................... | .................... | |
| M | 3608 hCoV-19/England/SHEF-C78FE/2020\|EPI_ISL_585030\|202 | .................... | .................... | .................... | .................... | .................... | .................... | |
| M | 3611 hCoV-19/England/SHEF-C907C/2020\|EPI_ISL_585034\|202 | .................... | .................... | .................... | .................... | .................... | .................... | |
| M | 3612 hCoV-19/Canada/ON-S1224/2020\|EPI_ISL_586300\|2020-0 | .................... | .................... | .................... | .................... | .................... | .................... | |
| M | 3613 hCoV-19/Canada/ON-S1225/2020\|EPI_ISL_586301\|2020-0 | .................... | .................... | .................... | .................... | .................... | .................... | |
| M | 3614 hCoV-19/Canada/ON-S1312/2020\|EPI_ISL_586349\|2020-0 | .................... | .................... | .................... | .................... | .................... | .................... | |
| M | 3615 hCoV-19/Canada/ON-S1313/2020\|EPI_ISL_586350\|2020-0 | .................... | .................... | .................... | .................... | .................... | .................... | |
| M | 3616 hCoV-19/Canada/ON-S153/2020\|EPI_ISL_586364\|2020-03 | .................... | .................... | .................... | .................... | .................... | .................... | |
| M | 3617 hCoV-19/England/QEUH-A212DC/2020\|EPI_ISL_587600\|20 | .................... | .................... | .................... | .................... | .................... | .................... | |
| M | 3618 hCoV-19/England/QEUH-A2243F/2020\|EPI_ISL_587601\|20 | .................... | .................... | .................... | .................... | .................... | .................... | |
| M | 3619 hCoV-19/England/QEUH-A1FCF1/2020\|EPI_ISL_587602\|20 | .................... | .................... | .................... | .................... | .................... | .................... | |

| | | | | | | | |
|---|---|---|---|---|---|---|---|
| M | 3620 hCoV-19/England/QEUH-A2045F/2020\|EPI_ISL_587603\|20 | ···················· | ···················· | ···················· | ···················· | ···················· | ···················· |
| M | 3621 hCoV-19/England/QEUH-A209E7/2020\|EPI_ISL_587604\|20 | ···················· | ···················· | ···················· | ···················· | ···················· | ···················· |
| M | 3622 hCoV-19/England/QEUH-A224B7/2020\|EPI_ISL_587605\|20 | ···················· | ···················· | ···················· | ···················· | ···················· | ···················· |
| M | 3623 hCoV-19/England/QEUH-A215A6/2020\|EPI_ISL_587606\|20 | ···················· | ···················· | ···················· | ···················· | ···················· | ···················· |
| M | 3624 hCoV-19/England/QEUH-A2096F/2020\|EPI_ISL_587608\|20 | ···················· | ···················· | ···················· | ···················· | ···················· | ···················· |
| M | 3625 hCoV-19/England/QEUH-A2244E/2020\|EPI_ISL_587609\|20 | ···················· | ···················· | ···················· | ···················· | ···················· | ···················· |
| M | 3626 hCoV-19/England/QEUH-A1F9FA/2020\|EPI_ISL_587610\|20 | ···················· | ···················· | ···················· | ···················· | ···················· | ···················· |
| M | 3627 hCoV-19/England/QEUH-A20352/2020\|EPI_ISL_587611\|20 | ···················· | ···················· | ···················· | ···················· | ···················· | ···················· |
| M | 3628 hCoV-19/England/QEUH-A202FB/2020\|EPI_ISL_587612\|20 | ···················· | ···················· | ···················· | ···················· | ···················· | ···················· |
| M | 3629 hCoV-19/England/QEUH-A1F9CD/2020\|EPI_ISL_587613\|20 | ···················· | ···················· | ···················· | ···················· | ···················· | ···················· |
| M | 3630 hCoV-19/England/QEUH-A20334/2020\|EPI_ISL_587614\|20 | ···················· | ···················· | ···················· | ···················· | ···················· | ···················· |
| M | 3631 hCoV-19/England/QEUH-A20246/2020\|EPI_ISL_587615\|20 | ···················· | ···················· | ···················· | ···················· | ···················· | ···················· |
| M | 3632 hCoV-19/England/QEUH-A2105A/2020\|EPI_ISL_587616\|20 | ···················· | ···················· | ···················· | ···················· | ···T················ | ···················· |
| M | 3633 hCoV-19/England/QEUH-A22709/2020\|EPI_ISL_587617\|20 | ···················· | ···················· | ···················· | ···················· | ···················· | ···················· |
| M | 3634 hCoV-19/England/QEUH-A21EC9/2020\|EPI_ISL_587618\|20 | ···················· | ···················· | ···················· | ···················· | ···················· | ···················· |
| M | 3635 hCoV-19/England/QEUH-A215E2/2020\|EPI_ISL_587619\|20 | ···················· | ···················· | ···················· | ···················· | ···················· | ···················· |
| M | 3636 hCoV-19/England/QEUH-A1FCD3/2020\|EPI_ISL_587620\|20 | ···················· | ···················· | ···················· | ···················· | ···················· | ···················· |
| M | 3637 hCoV-19/England/QEUH-A20237/2020\|EPI_ISL_587621\|20 | ···················· | ···················· | ···················· | ···················· | ···················· | ···················· |
| M | 3638 hCoV-19/England/QEUH-A2246C/2020\|EPI_ISL_587622\|20 | ···················· | ···················· | ···················· | ···················· | ···················· | ···················· |
| M | 3639 hCoV-19/England/QEUH-A224A8/2020\|EPI_ISL_587623\|20 | ···················· | ···················· | ···················· | ···················· | ···················· | ···················· |
| M | 3640 hCoV-19/England/QEUH-A20D54/2020\|EPI_ISL_587624\|20 | ···················· | ···················· | ···················· | ···················· | ···················· | ···················· |
| M | 3641 hCoV-19/England/QEUH-A210D2/2020\|EPI_ISL_587625\|20 | ···················· | ···················· | ···················· | ···················· | ·····T·············· | ···················· |
| M | 3642 hCoV-19/England/QEUH-A208EA/2020\|EPI_ISL_587626\|20 | ···················· | ···················· | ···················· | ···················· | ···················· | ···················· |
| M | 3643 hCoV-19/England/QEUH-A21078/2020\|EPI_ISL_587627\|20 | ···················· | ···················· | ···················· | ···················· | ···················· | ···················· |
| M | 3644 hCoV-19/England/QEUH-A21096/2020\|EPI_ISL_587628\|20 | ···················· | ···················· | ···················· | ···················· | ···················· | ···················· |
| M | 3645 hCoV-19/England/QEUH-A20756/2020\|EPI_ISL_587629\|20 | ···················· | ···················· | ···················· | ···················· | ···················· | ···················· |
| M | 3646 hCoV-19/England/QEUH-A21E6F/2020\|EPI_ISL_587631\|20 | ···················· | ···················· | ···················· | ···················· | ···················· | ···················· |
| M | 3647 hCoV-19/England/QEUH-A209AB/2020\|EPI_ISL_587632\|20 | ···················· | ···················· | ···················· | ···················· | ···················· | ···················· |
| M | 3648 hCoV-19/England/QEUH-A202A0/2020\|EPI_ISL_587677\|20 | ···················· | ···················· | ···················· | ···················· | ···················· | ···················· |
| M | 3649 hCoV-19/England/QEUH-A22499/2020\|EPI_ISL_587678\|20 | ···················· | ···················· | ···················· | ···················· | ···················· | ···················· |
| M | 3650 hCoV-19/England/QEUH-A21245/2020\|EPI_ISL_587679\|20 | ···················· | ·A·················· | ···················· | ···················· | ···················· | ···················· |
| M | 3651 hCoV-19/England/QEUH-A22226/2020\|EPI_ISL_587680\|20 | ···················· | ···················· | ···················· | ···················· | ···················· | ···················· |
| M | 3652 hCoV-19/England/QEUH-A1FE91/2020\|EPI_ISL_587681\|20 | ···················· | ·A·················· | ···················· | ···················· | ···················· | ··A················· |
| M | 3653 hCoV-19/England/QEUH-A209BA/2020\|EPI_ISL_587683\|20 | ···················· | ···················· | ···················· | ···················· | ···················· | ···················· |
| M | 3654 hCoV-19/England/QEUH-A20765/2020\|EPI_ISL_587684\|20 | ···················· | ···················· | ···················· | ···················· | ···················· | ···················· |
| M | 3655 hCoV-19/England/QEUH-A224E4/2020\|EPI_ISL_587685\|20 | ···················· | ···················· | ···················· | ···················· | ···················· | ···················· |
| M | 3656 hCoV-19/England/QEUH-A208AE/2020\|EPI_ISL_587686\|20 | ···················· | ···················· | ···················· | ···················· | ···················· | ···················· |
| M | 3657 hCoV-19/England/QEUH-A212CD/2020\|EPI_ISL_587687\|20 | ···················· | ···················· | ···················· | ···················· | ···················· | ···················· |
| M | 3658 hCoV-19/England/QEUH-A2054D/2020\|EPI_ISL_587688\|20 | ···················· | ···················· | ···················· | ···················· | ···T················ | ···················· |
| M | 3659 hCoV-19/England/QEUH-A20914/2020\|EPI_ISL_587689\|20 | ···················· | ···················· | ···················· | ···················· | ···················· | ···················· |
| M | 3660 hCoV-19/England/QEUH-A1FCA6/2020\|EPI_ISL_587690\|20 | ···················· | ···················· | ···················· | ···················· | ···················· | ···················· |
| M | 3661 hCoV-19/England/QEUH-A20361/2020\|EPI_ISL_587691\|20 | ···················· | ···················· | ···················· | ···················· | ···················· | ···················· |
| M | 3662 hCoV-19/England/QEUH-A21227/2020\|EPI_ISL_587693\|20 | ···················· | ···················· | ···················· | ···················· | ·····T·············· | ···················· |
| M | 3663 hCoV-19/England/QEUH-A208CC/2020\|EPI_ISL_587694\|20 | ···················· | ···················· | ···················· | ···················· | ···················· | ···················· |
| M | 3664 hCoV-19/England/QEUH-A221ED/2020\|EPI_ISL_587695\|20 | ···················· | ···················· | ···················· | ···················· | ···················· | ···················· |
| M | 3665 hCoV-19/England/QEUH-A1F9BE/2020\|EPI_ISL_587696\|20 | ···················· | ···················· | ···················· | ···················· | ···················· | ···················· |
| M | 3666 hCoV-19/England/QEUH-A2100F/2020\|EPI_ISL_587697\|20 | ···················· | ···················· | ···················· | ···················· | ···················· | ···················· |
| M | 3667 hCoV-19/England/QEUH-A21E7E/2020\|EPI_ISL_587698\|20 | ···················· | ···················· | ···················· | ···················· | ···················· | ···················· |
| M | 3668 hCoV-19/England/QEUH-9F56F4/2020\|EPI_ISL_588900\|20 | ···················· | ···················· | ···················· | ···················· | ···················· | ···················· |
| M | 3669 hCoV-19/England/QEUH-9F9371/2020\|EPI_ISL_588901\|20 | ···················· | ···················· | ···················· | ···················· | ···················· | ···················· |
| M | 3670 hCoV-19/England/QEUH-9F5973/2020\|EPI_ISL_588902\|20 | ···················· | ·A·················· | ···················· | ···················· | ···················· | ···················· |
| M | 3671 hCoV-19/England/QEUH-9F576A/2020\|EPI_ISL_588903\|20 | ···················· | ···················· | ···················· | ···················· | ···················· | ···················· |
| M | 3672 hCoV-19/England/QEUH-9F8C86/2020\|EPI_ISL_588904\|20 | ···················· | ···················· | ···················· | ···················· | ···················· | ···················· |
| M | 3673 hCoV-19/Wales/QEUH-9F5EED/2020\|EPI_ISL_588905\|2020 | ···················· | ···················· | ···················· | ···················· | ···················· | ···················· |
| M | 3674 hCoV-19/England/QEUH-9F8248/2020\|EPI_ISL_588906\|20 | ···················· | ···················· | ···················· | ···················· | ···················· | ···················· |
| M | 3675 hCoV-19/England/QEUH-9F6338/2020\|EPI_ISL_588907\|20 | ···················· | ·A·················· | ···················· | ···················· | ···················· | ··A················· |
| M | 3676 hCoV-19/England/QEUH-9F87B2/2020\|EPI_ISL_588908\|20 | ···················· | ···················· | ···················· | ···················· | ···················· | ···················· |
| M | 3677 hCoV-19/Wales/QEUH-9F6444/2020\|EPI_ISL_588909\|2020 | ···················· | ···················· | ···················· | ···················· | ···················· | ···················· |
| M | 3678 hCoV-19/England/QEUH-9F57D3/2020\|EPI_ISL_588910\|20 | ···················· | ···················· | ···················· | ···················· | ···················· | ···················· |
| M | 3679 hCoV-19/England/QEUH-9F862E/2020\|EPI_ISL_588911\|20 | ···················· | ···················· | ···················· | ···················· | ···················· | ···················· |
| M | 3680 hCoV-19/England/QEUH-9F8767/2020\|EPI_ISL_588913\|20 | ···················· | ···················· | ···················· | ···················· | ···················· | ···················· |
| M | 3681 hCoV-19/England/QEUH-9F85F4/2020\|EPI_ISL_588914\|20 | ···················· | ···················· | ···················· | ···················· | ···················· | ···················· |
| M | 3682 hCoV-19/England/QEUH-9F8D1A/2020\|EPI_ISL_588915\|20 | ···················· | ···················· | ···················· | ···················· | ···················· | ···················· |
| M | 3683 hCoV-19/England/QEUH-9F62E0/2020\|EPI_ISL_588916\|20 | ···················· | ···················· | ···················· | ···················· | ···················· | ···················· |
| M | 3684 hCoV-19/England/QEUH-9F5964/2020\|EPI_ISL_588917\|20 | ···················· | ···················· | ···················· | ···················· | ···················· | ···················· |
| M | 3685 hCoV-19/England/QEUH-9F6347/2020\|EPI_ISL_588918\|20 | ···················· | ···················· | ···················· | ···················· | ···················· | ···················· |
| M | 3686 hCoV-19/England/QEUH-9F814B/2020\|EPI_ISL_588920\|20 | ···················· | ···················· | ···················· | ···················· | ···················· | ···················· |
| M | 3687 hCoV-19/England/QEUH-9F9308/2020\|EPI_ISL_588921\|20 | ···················· | ···················· | ···················· | ···················· | ···················· | ···················· |
| M | 3688 hCoV-19/England/QEUH-9F9089/2020\|EPI_ISL_588922\|20 | ···················· | ···················· | ···················· | ···················· | ···················· | ···················· |
| M | 3689 hCoV-19/England/QEUH-9F574C/2020\|EPI_ISL_588923\|20 | ···················· | ···················· | ···················· | ···················· | ···················· | ···················· |
| M | 3690 hCoV-19/England/QEUH-9F621D/2020\|EPI_ISL_588924\|20 | ···················· | ···················· | ···················· | ···················· | ···················· | ···················· |
| M | 3691 hCoV-19/England/QEUH-9F7CC3/2020\|EPI_ISL_588925\|20 | ···················· | ···················· | ···················· | ···················· | ·····T·············· | ···················· |
| M | 3692 hCoV-19/England/QEUH-9F8C3B/2020\|EPI_ISL_588926\|20 | ···················· | ···················· | ···················· | ···················· | ···················· | ···················· |
| M | 3693 hCoV-19/England/QEUH-9F8E53/2020\|EPI_ISL_588928\|20 | ···················· | ···················· | ···················· | ···················· | ···················· | ···················· |
| M | 3694 hCoV-19/England/QEUH-9F89E9/2020\|EPI_ISL_588929\|20 | ···················· | ·A·················· | ···················· | ···················· | ···················· | ··A················· |
| M | 3695 hCoV-19/England/QEUH-9F5A43/2020\|EPI_ISL_588930\|20 | ···················· | ···················· | ···················· | ···················· | ···················· | ···················· |
| M | 3696 hCoV-19/England/QEUH-9F60C8/2020\|EPI_ISL_588931\|20 | ···················· | ···················· | ···················· | ···················· | ···················· | ···················· |
| M | 3697 hCoV-19/England/QEUH-9F7D0C/2020\|EPI_ISL_588932\|20 | ···················· | ···················· | ···················· | ···················· | ···················· | ···················· |
| M | 3698 hCoV-19/England/QEUH-9F5A25/2020\|EPI_ISL_588933\|20 | ···················· | ···················· | ···················· | ···················· | ···················· | ···················· |
| M | 3699 hCoV-19/England/QEUH-9F5A07/2020\|EPI_ISL_588934\|20 | ···················· | ···················· | ···················· | ···················· | ···················· | ···················· |

| | | | | | | | |
|---|---|---|---|---|---|---|---|
| M | 3700 hCoV-19/England/QEUH-9F86B5/2020\|EPI_ISL_588935\|20 | ···················· | ···················· | ···················· | ···················· | ····T··············· | ···················· |
| M | 3701 hCoV-19/England/QEUH-9F899E/2020\|EPI_ISL_588936\|20 | ···················· | ···················· | ···················· | ···················· | ···················· | ···················· |
| M | 3702 hCoV-19/England/QEUH-9F9423/2020\|EPI_ISL_588937\|20 | ···················· | ···················· | ···················· | ···················· | ···················· | ···················· |
| M | 3703 hCoV-19/England/QEUH-9F8828/2020\|EPI_ISL_588938\|20 | ···················· | ···················· | ···················· | ···················· | ···················· | ···················· |
| M | 3704 hCoV-19/England/QEUH-9F5A70/2020\|EPI_ISL_588939\|20 | ···················· | ···················· | ···················· | ···················· | ···················· | ···················· |
| M | 3705 hCoV-19/England/QEUH-9F60AA/2020\|EPI_ISL_588941\|20 | ···················· | ···················· | ···················· | ···················· | ···················· | ···················· |
| M | 3706 hCoV-19/England/QEUH-9F8A13/2020\|EPI_ISL_588942\|20 | ···················· | ·A·················· | ···················· | ···················· | ···················· | ··A················· |
| M | 3707 hCoV-19/England/QEUH-9F93F9/2020\|EPI_ISL_588943\|20 | ···················· | ···················· | ···················· | ···················· | ···················· | ···················· |
| M | 3708 hCoV-19/England/QEUH-9F8C68/2020\|EPI_ISL_588944\|20 | ···················· | ···················· | ···················· | ···················· | ···················· | ···················· |
| M | 3709 hCoV-19/England/QEUH-9F8C77/2020\|EPI_ISL_588945\|20 | ···················· | ···················· | ···················· | ···················· | ···················· | ···················· |
| M | 3710 hCoV-19/England/QEUH-9F8460/2020\|EPI_ISL_588946\|20 | ···················· | ···················· | ···················· | ···················· | ···················· | ···················· |
| M | 3711 hCoV-19/England/QEUH-9F5991/2020\|EPI_ISL_588947\|20 | ···················· | ···················· | ···················· | ···················· | ···················· | ···················· |
| M | 3712 hCoV-19/England/QEUH-9F5779/2020\|EPI_ISL_588948\|20 | ···················· | ···················· | ···················· | ···················· | ···················· | ···················· |
| M | 3713 hCoV-19/England/QEUH-9F8372/2020\|EPI_ISL_588949\|20 | ···················· | ···················· | ···················· | ···················· | ···················· | ···················· |
| M | 3714 hCoV-19/England/QEUH-9F8D0B/2020\|EPI_ISL_588950\|20 | ···················· | ···················· | ···················· | ···················· | ···················· | ···················· |
| M | 3715 hCoV-19/England/QEUH-9F63CF/2020\|EPI_ISL_588951\|20 | ···················· | ···················· | ···················· | ···················· | ···················· | ···················· |
| M | 3716 hCoV-19/England/QEUH-9F5955/2020\|EPI_ISL_588952\|20 | ···················· | ···················· | ···················· | ···················· | ···················· | ···················· |
| M | 3717 hCoV-19/England/QEUH-9F8D29/2020\|EPI_ISL_588953\|20 | ···················· | ···················· | ···················· | ···················· | ···················· | ···················· |
| M | 3718 hCoV-19/England/QEUH-9F9229/2020\|EPI_ISL_588954\|20 | ···················· | ···················· | ···················· | ···················· | ···················· | ···················· |
| M | 3719 hCoV-19/England/QEUH-9F821B/2020\|EPI_ISL_588955\|20 | ···················· | ···················· | ···················· | ···················· | ···················· | ···················· |
| M | 3720 hCoV-19/England/QEUH-9F93EA/2020\|EPI_ISL_588956\|20 | ···················· | ···················· | ···················· | ···················· | ···················· | ···················· |
| M | 3721 hCoV-19/England/QEUH-9F8D47/2020\|EPI_ISL_588957\|20 | ···················· | ···················· | ···················· | ···················· | ···················· | ···················· |
| M | 3722 hCoV-19/England/QEUH-9F8785/2020\|EPI_ISL_588959\|20 | ···················· | ···················· | ···················· | ···················· | ···················· | ···················· |
| M | 3723 hCoV-19/England/QEUH-9F84D9/2020\|EPI_ISL_588960\|20 | ···················· | ···················· | ···················· | ···················· | ···················· | ···················· |
| M | 3724 hCoV-19/England/QEUH-9F8C2C/2020\|EPI_ISL_588961\|20 | ···················· | ·A·················· | ···················· | ···················· | ···················· | ···················· |
| M | 3725 hCoV-19/England/QEUH-9F89F8/2020\|EPI_ISL_588963\|20 | ···················· | ···················· | ···················· | ···················· | ···················· | ···················· |
| M | 3726 hCoV-19/Australia/VIC12997/2020\|EPI_ISL_592143\|202 | ···················· | ···················· | ···················· | ···················· | ···················· | ···················· |
| M | 3727 hCoV-19/Australia/VIC12999/2020\|EPI_ISL_592144\|202 | ···················· | ···················· | ···················· | ···················· | ···················· | ···················· |
| M | 3728 hCoV-19/Australia/VIC13000/2020\|EPI_ISL_592145\|202 | ···················· | ···················· | ···················· | ···················· | ···················· | ···················· |
| M | 3729 hCoV-19/Australia/VIC13003/2020\|EPI_ISL_592146\|202 | ···················· | ···················· | ···················· | ···················· | ···················· | ···················· |
| M | 3730 hCoV-19/Australia/VIC13008/2020\|EPI_ISL_592147\|202 | ···················· | ···················· | ···················· | ···················· | ···················· | ···················· |
| M | 3731 hCoV-19/Australia/VIC13012/2020\|EPI_ISL_592148\|202 | ···················· | ···················· | ···················· | ···················· | ···················· | ···················· |
| M | 3732 hCoV-19/Australia/VIC13013/2020\|EPI_ISL_592149\|202 | ···················· | ···················· | ···················· | ···················· | ···················· | ···················· |
| M | 3733 hCoV-19/Australia/VIC13014/2020\|EPI_ISL_592150\|202 | ···················· | ···················· | ···················· | ···················· | ···················· | ···················· |
| M | 3734 hCoV-19/Australia/VIC13019/2020\|EPI_ISL_592151\|202 | ···················· | ···················· | ···················· | ···················· | ···················· | ···················· |
| M | 3735 hCoV-19/Australia/VIC13020/2020\|EPI_ISL_592152\|202 | ···················· | ···················· | ···················· | ···················· | ···················· | ···················· |
| M | 3736 hCoV-19/Australia/VIC13021/2020\|EPI_ISL_592153\|202 | ···················· | ···················· | ··K················· | ···················· | ···················· | ···················· |
| M | 3737 hCoV-19/Australia/VIC13024/2020\|EPI_ISL_592154\|202 | ···················· | ···················· | ···················· | ···················· | ···················· | ···················· |
| M | 3738 hCoV-19/Australia/VIC13030/2020\|EPI_ISL_592155\|202 | ···················· | ···················· | ··K················· | ···················· | ···················· | ···················· |
| M | 3739 hCoV-19/Australia/VIC13031/2020\|EPI_ISL_592156\|202 | ···················· | ···················· | ···················· | ···················· | ···················· | ···················· |
| M | 3740 hCoV-19/Australia/VIC13033/2020\|EPI_ISL_592157\|202 | ···················· | ···················· | ···················· | ···················· | ···················· | ···················· |
| M | 3741 hCoV-19/Australia/VIC13034/2020\|EPI_ISL_592158\|202 | ···················· | ···················· | ··K················· | ···················· | ···················· | ···················· |
| M | 3742 hCoV-19/Australia/VIC13035/2020\|EPI_ISL_592159\|202 | ···················· | ···················· | ···················· | ···················· | ···················· | ···················· |
| M | 3743 hCoV-19/Australia/VIC13036/2020\|EPI_ISL_592160\|202 | ···················· | ···················· | ···················· | ···················· | ···················· | ···················· |
| M | 3744 hCoV-19/Australia/VIC13039/2020\|EPI_ISL_592161\|202 | ···················· | ···················· | ···················· | ···················· | ···················· | ···················· |
| M | 3745 hCoV-19/Australia/VIC13043/2020\|EPI_ISL_592162\|202 | ···················· | ···················· | ···················· | ···················· | ···················· | ···················· |
| M | 3746 hCoV-19/Australia/VIC13044/2020\|EPI_ISL_592163\|202 | ···················· | ···················· | ···················· | ···················· | ···················· | ···················· |
| M | 3747 hCoV-19/Australia/VIC13049/2020\|EPI_ISL_592164\|202 | ···················· | ···················· | ···················· | ···················· | ···················· | ···················· |
| M | 3748 hCoV-19/Australia/VIC13050/2020\|EPI_ISL_592165\|202 | ···················· | ···················· | ···················· | ···················· | ···················· | ···················· |
| M | 3749 hCoV-19/Australia/VIC13051/2020\|EPI_ISL_592166\|202 | ···················· | ···················· | ···················· | ···················· | ···················· | ···················· |
| M | 3750 hCoV-19/Australia/VIC13053/2020\|EPI_ISL_592167\|202 | ···················· | ···················· | ··K················· | ···················· | ···················· | ···················· |
| M | 3751 hCoV-19/Australia/VIC13054/2020\|EPI_ISL_592168\|202 | ···················· | ···················· | ···················· | ···················· | ···················· | ···················· |
| M | 3752 hCoV-19/Australia/VIC13055/2020\|EPI_ISL_592169\|202 | ···················· | ···················· | ···················· | ···················· | ···················· | ···················· |
| M | 3753 hCoV-19/Australia/VIC13061/2020\|EPI_ISL_592170\|202 | ···················· | ···················· | ···················· | ···················· | ···················· | ···················· |
| M | 3754 hCoV-19/Australia/VIC13063/2020\|EPI_ISL_592171\|202 | ···················· | ···················· | ···················· | ···················· | ···················· | ···················· |
| M | 3755 hCoV-19/Australia/VIC13064/2020\|EPI_ISL_592172\|202 | ···················· | ···················· | ··K················· | ···················· | ···················· | ···················· |
| M | 3756 hCoV-19/Australia/VIC13065/2020\|EPI_ISL_592173\|202 | ···················· | ···················· | ··N················· | ···················· | ···················· | ···················· |
| M | 3757 hCoV-19/Australia/VIC13067/2020\|EPI_ISL_592174\|202 | ···················· | ···················· | ··K················· | ···················· | ···················· | ···················· |
| M | 3758 hCoV-19/Australia/VIC13069/2020\|EPI_ISL_592175\|202 | ···················· | ···················· | ···················· | ···················· | ···················· | ···················· |
| M | 3759 hCoV-19/Australia/VIC13070/2020\|EPI_ISL_592176\|202 | ···················· | ···················· | ··K················· | ···················· | ···················· | ···················· |
| M | 3760 hCoV-19/Australia/VIC13073/2020\|EPI_ISL_592177\|202 | ···················· | ···················· | ··K················· | ···················· | ···················· | ···················· |
| M | 3761 hCoV-19/Australia/VIC13074/2020\|EPI_ISL_592178\|202 | ···················· | ···················· | ··K················· | ···················· | ···················· | ···················· |
| M | 3762 hCoV-19/Australia/VIC13076/2020\|EPI_ISL_592179\|202 | ···················· | ···················· | ··K················· | ···················· | ···················· | ···················· |
| M | 3763 hCoV-19/Australia/VIC13078/2020\|EPI_ISL_592180\|202 | ···················· | ···················· | ···················· | ···················· | ···················· | ···················· |
| M | 3764 hCoV-19/Australia/VIC13080/2020\|EPI_ISL_592181\|202 | ···················· | ···················· | ··K················· | ···················· | ···················· | ···················· |
| M | 3765 hCoV-19/Australia/VIC13083/2020\|EPI_ISL_592182\|202 | ···················· | ···················· | ··K················· | ···················· | ···················· | ···················· |
| M | 3766 hCoV-19/Australia/VIC13090/2020\|EPI_ISL_592183\|202 | ···················· | ···················· | ··K················· | ···················· | ···················· | ···················· |
| M | 3767 hCoV-19/Australia/VIC13091/2020\|EPI_ISL_592184\|202 | ···················· | ···················· | ···················· | ···················· | ···················· | ···················· |
| M | 3768 hCoV-19/Australia/VIC13092/2020\|EPI_ISL_592185\|202 | ···················· | ···················· | ··K················· | ···················· | ···················· | ···················· |
| M | 3769 hCoV-19/Australia/VIC13094/2020\|EPI_ISL_592186\|202 | ···················· | ···················· | ··K················· | ···················· | ············K······· | ···················· |
| M | 3770 hCoV-19/Australia/VIC13097/2020\|EPI_ISL_592187\|202 | ···················· | ···················· | ··K················· | ···················· | ···················· | ·RR················· |
| M | 3771 hCoV-19/Australia/VIC13100/2020\|EPI_ISL_592188\|202 | ···················· | ···················· | ···················· | ···················· | ···················· | ···················· |
| M | 3772 hCoV-19/Australia/VIC13115/2020\|EPI_ISL_592189\|202 | ···················· | ···················· | ···················· | ···················· | ···················· | ···················· |
| M | 3773 hCoV-19/Australia/VIC13119/2020\|EPI_ISL_592190\|202 | ···················· | ···················· | ··K················· | ···················· | ···················· | ···················· |
| M | 3774 hCoV-19/Australia/VIC13123/2020\|EPI_ISL_592191\|202 | ···················· | ···················· | ···················· | ···················· | ···················· | ···················· |
| M | 3775 hCoV-19/Australia/VIC13125/2020\|EPI_ISL_592192\|202 | ···················· | ···················· | ··K················· | ···················· | ···················· | ···················· |
| M | 3776 hCoV-19/Australia/VIC13133/2020\|EPI_ISL_592193\|202 | ···················· | ···················· | ···················· | ···················· | ···················· | ···················· |
| M | 3777 hCoV-19/Australia/VIC13135/2020\|EPI_ISL_592194\|202 | ···················· | ···················· | ···················· | ···················· | ···················· | ···················· |
| M | 3778 hCoV-19/Australia/VIC13137/2020\|EPI_ISL_592195\|202 | ···················· | ···················· | ···················· | ···················· | ···················· | ···················· |
| M | 3779 hCoV-19/Australia/VIC13147/2020\|EPI_ISL_592196\|202 | ···················· | ···················· | ···················· | ···················· | ···················· | ···················· |

| | | | | | | | | |
|---|---|---|---|---|---|---|---|---|
| M | 3780 | hCoV-19/Australia/VIC13152/2020\|EPI_ISL_592197\|202 | ···················· | ···················· | ···················· | ···················· | ···················· | ···················· |
| M | 3781 | hCoV-19/Australia/VIC13154/2020\|EPI_ISL_592198\|202 | ···················· | ···················· | ···················· | ···················· | ···················· | ···················· |
| M | 3782 | hCoV-19/Australia/VIC13167/2020\|EPI_ISL_592199\|202 | ···················· | ···················· | ···················· | ···················· | ···················· | ···················· |
| M | 3783 | hCoV-19/Australia/VIC15422/2020\|EPI_ISL_593300\|202 | ···················· | ···················· | ···················· | ···················· | ···················· | ···················· |
| M | 3784 | hCoV-19/Australia/VIC15424/2020\|EPI_ISL_593301\|202 | ···················· | ···················· | ···················· | ···················· | ···················· | ···················· |
| M | 3785 | hCoV-19/Australia/VIC15429/2020\|EPI_ISL_593302\|202 | ···················· | ···················· | ···················· | ···················· | ···················· | ···················· |
| M | 3786 | hCoV-19/Australia/VIC15430/2020\|EPI_ISL_593303\|202 | ···················· | ···················· | ···················· | ···················· | ···················· | ···················· |
| M | 3787 | hCoV-19/Australia/VIC15437/2020\|EPI_ISL_593304\|202 | ···················· | ···················· | ···················· | ···················· | ···················· | ···················· |
| M | 3788 | hCoV-19/Australia/VIC15438/2020\|EPI_ISL_593305\|202 | ···················· | ···················· | ···················· | ···················· | ···················· | ···················· |
| M | 3789 | hCoV-19/Australia/VIC15440/2020\|EPI_ISL_593306\|202 | ···················· | ···················· | ···················· | ···················· | ···················· | ···················· |
| M | 3790 | hCoV-19/Australia/VIC15444/2020\|EPI_ISL_593308\|202 | ···················· | ···················· | ···················· | ···················· | ···················· | ···················· |
| M | 3791 | hCoV-19/Australia/VIC15446/2020\|EPI_ISL_593309\|202 | ···················· | ···················· | ···················· | ···················· | ···················· | ···················· |
| M | 3793 | hCoV-19/Australia/VIC15842/2020\|EPI_ISL_593475\|202 | ···················· | ···················· | ···················· | ···················· | ···················· | ···················· |
| M | 3795 | hCoV-19/Australia/VIC3605/2020\|EPI_ISL_593477\|2020 | ···················· | ···················· | ···················· | ···················· | ···················· | ···················· |
| M | 3796 | hCoV-19/USA/MA-JLL-D18/2020\|EPI_ISL_593478\|2020-04 | ···················· | ···················· | ···················· | ···················· | ···················· | ···················· |
| M | 3797 | hCoV-19/USA/MA-JLL-D25/2020\|EPI_ISL_593479\|2020-05 | ···················· | ···················· | ···················· | ···················· | ···················· | ···················· |
| M | 3798 | hCoV-19/USA/MA-JLL-D75/2020\|EPI_ISL_593480\|2020-06 | ···················· | ···················· | ···················· | ···················· | ···················· | ···················· |
| M | 3799 | hCoV-19/Canada/ON-E1/2020\|EPI_ISL_593481\|2020-04-1 | ···················· | ···················· | ···················· | ···················· | ···················· | ···················· |
| M | 3800 | hCoV-19/Canada/ON-E7/2020\|EPI_ISL_593487\|2020-04-2 | ···················· | ···················· | ···················· | ···················· | ···················· | ···················· |
| M | 3801 | hCoV-19/Canada/ON-E9/2020\|EPI_ISL_593489\|2020-04-2 | ···················· | ···················· | ···················· | ···················· | ···················· | ···················· |
| M | 3802 | hCoV-19/Canada/ON-E10/2020\|EPI_ISL_593490\|2020-04- | ···················· | ···················· | ···················· | ···················· | ···················· | ···················· |
| M | 3803 | hCoV-19/Canada/ON-E11/2020\|EPI_ISL_593491\|2020-05- | ········Y··········· | ···················· | ···················· | ···················· | ···················· | ···················· |
| M | 3804 | hCoV-19/Canada/ON-E12/2020\|EPI_ISL_593492\|2020-05- | ···················· | ···················· | ···················· | ···················· | ···················· | ···················· |
| M | 3805 | hCoV-19/Canada/ON-E13/2020\|EPI_ISL_593493\|2020-05- | ···················· | ···················· | ···················· | ···················· | ···················· | ···················· |
| M | 3806 | hCoV-19/Canada/ON-E14/2020\|EPI_ISL_593494\|2020-05- | ···················· | ···················· | ···················· | ···················· | ···················· | ···················· |
| M | 3807 | hCoV-19/Canada/ON-E15/2020\|EPI_ISL_593495\|2020-05- | ···················· | ···················· | ···················· | ···················· | ···················· | ···················· |
| M | 3808 | hCoV-19/Canada/ON-E16/2020\|EPI_ISL_593496\|2020-05- | ···················· | ···················· | ···················· | ···················· | ···················· | ···················· |
| M | 3809 | hCoV-19/Canada/ON-E18/2020\|EPI_ISL_593497\|2020-05- | ···················· | ···················· | ···················· | ···················· | ···················· | ···················· |
| M | 3810 | hCoV-19/Canada/ON-E19/2020\|EPI_ISL_593498\|2020-05- | ···················· | ···················· | ···················· | ···················· | ···················· | ···················· |
| M | 3811 | hCoV-19/Canada/ON-E20/2020\|EPI_ISL_593499\|2020-05- | ···················· | ···················· | ···················· | ···················· | ···················· | ···················· |
| M | 3812 | hCoV-19/England/CAMC-9EC818/2020\|EPI_ISL_594620\|20 | ···················· | ···················· | ···················· | ···················· | ···················· | ···················· |
| M | 3813 | hCoV-19/England/CAMC-9EC845/2020\|EPI_ISL_594622\|20 | ···················· | ···················· | ···················· | ···················· | ···················· | ···················· |
| M | 3814 | hCoV-19/England/CAMC-9EC863/2020\|EPI_ISL_594623\|20 | ···················· | ···················· | ···················· | ···················· | ···················· | ···················· |
| M | 3815 | hCoV-19/England/CAMC-9EC872/2020\|EPI_ISL_594624\|20 | ···················· | ···················· | ···················· | ···················· | ···················· | ···················· |
| M | 3816 | hCoV-19/England/CAMC-9EC881/2020\|EPI_ISL_594625\|20 | ···················· | ···················· | ···················· | ···················· | ···················· | ···················· |
| M | 3817 | hCoV-19/England/CAMC-9EC890/2020\|EPI_ISL_594626\|20 | ···················· | ···················· | ···················· | ···················· | ···················· | ···················· |
| M | 3818 | hCoV-19/England/CAMC-9EC8FA/2020\|EPI_ISL_594627\|20 | ···················· | ···················· | ···················· | ···················· | ······T············· | ···················· |
| M | 3819 | hCoV-19/England/CAMC-9EC906/2020\|EPI_ISL_594628\|20 | ···················· | ···················· | ···················· | ···················· | ···················· | ···················· |
| M | 3820 | hCoV-19/England/CAMC-9EC915/2020\|EPI_ISL_594629\|20 | ···················· | ···················· | ···················· | ···················· | ···················· | ···················· |
| M | 3821 | hCoV-19/England/CAMC-9EC951/2020\|EPI_ISL_594630\|20 | ···················· | ···················· | ···················· | ···················· | ···················· | ···················· |
| M | 3822 | hCoV-19/England/CAMC-9EC97F/2020\|EPI_ISL_594631\|20 | ···················· | ···················· | ···················· | ···················· | ······T············· | ···················· |
| M | 3823 | hCoV-19/England/CAMC-9EC99D/2020\|EPI_ISL_594632\|20 | ···················· | ···················· | ···················· | ···················· | ···················· | ···················· |
| M | 3824 | hCoV-19/England/CAMC-9EC9BB/2020\|EPI_ISL_594633\|20 | ···················· | ···················· | ···················· | ···················· | ···················· | ···················· |
| M | 3825 | hCoV-19/England/CAMC-9EC9F7/2020\|EPI_ISL_594634\|20 | ···················· | ···················· | ···················· | ···················· | ···················· | ···················· |
| M | 3826 | hCoV-19/England/CAMC-9ECA21/2020\|EPI_ISL_594635\|20 | ···················· | ···················· | ···················· | ···················· | ···················· | ···················· |
| M | 3827 | hCoV-19/England/CAMC-9ECA7C/2020\|EPI_ISL_594636\|20 | ···················· | ·A·················· | ···················· | ···················· | ···················· | ··A················· |
| M | 3828 | hCoV-19/England/CAMC-9ECAD6/2020\|EPI_ISL_594639\|20 | ···················· | ···················· | ···················· | ···················· | ···················· | ···················· |
| M | 3829 | hCoV-19/Scotland/QEUH-A4905C/2020\|EPI_ISL_595950\|2 | ···················· | ···················· | ···················· | ···················· | ···················· | ···················· |
| M | 3831 | hCoV-19/Scotland/QEUH-A490B6/2020\|EPI_ISL_595954\|2 | ···················· | ···················· | ···················· | ···················· | ···················· | ···················· |
| M | 3832 | hCoV-19/England/QEUH-A492CF/2020\|EPI_ISL_595956\|20 | ···················· | ···················· | ···················· | ···················· | ···················· | ···················· |
| M | 3833 | hCoV-19/Scotland/QEUH-A492DE/2020\|EPI_ISL_595957\|2 | ···················· | ···················· | ···················· | ···················· | ···················· | ···················· |
| M | 3834 | hCoV-19/Scotland/QEUH-A49423/2020\|EPI_ISL_595960\|2 | ···················· | ···················· | ···················· | ···················· | ···················· | ···················· |
| M | 3835 | hCoV-19/England/QEUH-A49B88/2020\|EPI_ISL_595962\|20 | ···················· | ···················· | ···················· | ···················· | ···················· | ···················· |
| M | 3836 | hCoV-19/England/QEUH-A49B97/2020\|EPI_ISL_595963\|20 | ···················· | ···················· | ···················· | ···················· | ···················· | ···················· |
| M | 3837 | hCoV-19/Scotland/QEUH-A4AA20/2020\|EPI_ISL_595965\|2 | ···················· | ···················· | ···················· | ···················· | ···················· | ···················· |
| M | 3838 | hCoV-19/Scotland/QEUH-A4AA6C/2020\|EPI_ISL_595967\|2 | ···················· | ···················· | ···················· | ···················· | ···················· | ···················· |
| M | 3839 | hCoV-19/Scotland/QEUH-A4AAB7/2020\|EPI_ISL_595969\|2 | ···················· | ···················· | ···················· | ···················· | ···················· | ···················· |
| M | 3841 | hCoV-19/Singapore/1065/2020\|EPI_ISL_583900\|2020-08 | ···················· | ···················· | ···················· | ···················· | ···················· | ···················· |
| M | 3842 | hCoV-19/Italy/TUS-DBM-2/2020\|EPI_ISL_583953\|2020-0 | ···················· | ···················· | ···················· | ···················· | ···················· | ···················· |
| M | 3843 | hCoV-19/Italy/TUS-DBM-10/2020\|EPI_ISL_583954\|2020- | ···················· | ···················· | ···················· | ···················· | ···················· | ···················· |
| M | 3844 | hCoV-19/Italy/TUS-DBM-11/2020\|EPI_ISL_583955\|2020- | ···················· | ···················· | ···················· | ···················· | ···················· | ···················· |
| M | 3845 | hCoV-19/Italy/TUS-DBM-18/2020\|EPI_ISL_583956\|2020- | ···················· | ···················· | ···················· | ···················· | ···················· | ···················· |
| M | 3846 | hCoV-19/Italy/TUS-DBM-19/2020\|EPI_ISL_583957\|2020- | ···················· | ···················· | ···················· | ···················· | ···················· | ···················· |
| M | 3847 | hCoV-19/Italy/TUS-DBM-21/2020\|EPI_ISL_583958\|2020- | ···················· | ···················· | ···················· | ···················· | ···················· | ···················· |
| M | 3848 | hCoV-19/Italy/TUS-DBM-4/2020\|EPI_ISL_583959\|2020-0 | ···················· | ···················· | ···················· | ···················· | ···················· | ···················· |
| M | 3849 | hCoV-19/Italy/TUS-DBM-6/2020\|EPI_ISL_583960\|2020-0 | ···················· | ···················· | ···················· | ···················· | ···················· | ···················· |
| M | 3850 | hCoV-19/Italy/TUS-DBM-7/2020\|EPI_ISL_583961\|2020-0 | ···················· | ···················· | ···················· | ···················· | ···················· | ···················· |
| M | 3851 | hCoV-19/Italy/TUS-DBM-8/2020\|EPI_ISL_583962\|2020-0 | ···················· | NN·················· | ···················· | ···················· | ···················· | ···················· |
| M | 3852 | hCoV-19/England/204180123/2020\|EPI_ISL_583968\|2020 | ···················· | ···················· | ···················· | ···················· | ···················· | ···················· |
| M | 3853 | hCoV-19/England/204180124/2020\|EPI_ISL_583969\|2020 | ···················· | ···················· | ···················· | ···················· | ···················· | ···················· |
| M | 3854 | hCoV-19/England/204180261/2020\|EPI_ISL_583992\|2020 | ···················· | ···················· | ···················· | ···················· | ···················· | ···················· |
| M | 3855 | hCoV-19/England/204180262/2020\|EPI_ISL_583993\|2020 | ···················· | ···················· | ···················· | ···················· | ···················· | ···················· |
| M | 3856 | hCoV-19/USA/DE-DHSS-F918022/2020\|EPI_ISL_583994\|20 | ···················· | ···················· | ···················· | ···················· | ···················· | ···················· |
| M | 3857 | hCoV-19/Wales/PHWC-47D495/2020\|EPI_ISL_586148\|2020 | ···················· | ···················· | ···················· | ···················· | ···················· | ···················· |
| M | 3858 | hCoV-19/Wales/PHWC-47D4A4/2020\|EPI_ISL_586149\|2020 | ···················· | ···················· | ···················· | ···················· | ···················· | ···················· |
| M | 3859 | hCoV-19/Wales/PHWC-47D4B3/2020\|EPI_ISL_586150\|2020 | ···················· | ···················· | ···················· | ···················· | ···················· | ···················· |
| M | 3860 | hCoV-19/Wales/PHWC-47D4C2/2020\|EPI_ISL_586151\|2020 | ···················· | ···················· | ···················· | ···················· | ···················· | ···················· |
| M | 3863 | hCoV-19/Wales/PHWC-47D538/2020\|EPI_ISL_586157\|2020 | ···················· | ···················· | ···················· | ···················· | ···················· | ···················· |
| M | 3865 | hCoV-19/Wales/PHWC-47D583/2020\|EPI_ISL_586161\|2020 | ···················· | ···················· | ···················· | ···················· | ···················· | ···················· |
| M | 3866 | hCoV-19/Wales/PHWC-47D5DE/2020\|EPI_ISL_586166\|2020 | ···················· | ···················· | ···················· | ···················· | ···················· | ···················· |

| | | | | | | | | |
|---|---|---|---|---|---|---|---|---|
| M | 3869 hCoV-19/Wales/PHWC-47D626/2020\|EPI_ISL_586169\|2020 | ···················· | ···················· | ···················· | ···················· | NNN················· | ···················· | 1 |
| M | 3870 hCoV-19/Wales/PHWC-47D635/2020\|EPI_ISL_586170\|2020 | ···················· | ···················· | ···················· | ···················· | ···················· | ···················· | |
| M | 3872 hCoV-19/Wales/PHWC-47D662/2020\|EPI_ISL_586173\|2020 | ···················· | ···················· | ···················· | ···················· | ···················· | ···················· | |
| M | 3873 hCoV-19/Wales/PHWC-47D671/2020\|EPI_ISL_586174\|2020 | ···················· | ···················· | ···················· | ···················· | ···················· | ···················· | |
| M | 3874 hCoV-19/Wales/PHWC-47D680/2020\|EPI_ISL_586175\|2020 | ···················· | ···················· | ···················· | ···················· | ···················· | ···················· | |
| M | 3877 hCoV-19/Wales/PHWC-47D6CC/2020\|EPI_ISL_586179\|2020 | ···················· | ···················· | ···················· | ···················· | NNN················· | ···················· | 1 |
| M | 3879 hCoV-19/Wales/PHWC-47D6EA/2020\|EPI_ISL_586181\|2020 | ···················· | ···················· | ···················· | ···················· | ···················· | ···················· | |
| M | 3880 hCoV-19/Wales/PHWC-47D705/2020\|EPI_ISL_586183\|2020 | ···················· | ···················· | ···················· | ···················· | ···················· | ···················· | |
| M | 3881 hCoV-19/Wales/PHWC-47D714/2020\|EPI_ISL_586184\|2020 | ···················· | ···················· | ···················· | ···················· | NNN················· | ···················· | 1 |
| M | 3882 hCoV-19/Wales/PHWC-47D732/2020\|EPI_ISL_586185\|2020 | ···················· | ···················· | ···················· | ···················· | ···················· | ···················· | |
| M | 3883 hCoV-19/Wales/PHWC-47D741/2020\|EPI_ISL_586186\|2020 | ···················· | ···················· | ···················· | ···················· | ···················· | ···················· | |
| M | 3884 hCoV-19/Wales/PHWC-47D750/2020\|EPI_ISL_586187\|2020 | ···················· | ···················· | ···················· | ···················· | ···················· | ···················· | |
| M | 3885 hCoV-19/Wales/PHWC-47D78D/2020\|EPI_ISL_586189\|2020 | ···················· | ···················· | ···················· | ···················· | ···················· | ···················· | |
| M | 3888 hCoV-19/Wales/PHWC-47D7BA/2020\|EPI_ISL_586192\|2020 | ···················· | ···················· | ···················· | ···················· | ···················· | ···················· | |
| M | 3889 hCoV-19/Wales/PHWC-47D7F6/2020\|EPI_ISL_586195\|2020 | ···················· | ···················· | ···················· | ···················· | NNN················· | ···················· | 1 |
| M | 3890 hCoV-19/Wales/PHWC-47D802/2020\|EPI_ISL_586196\|2020 | ···················· | ···················· | ···················· | ···················· | ···················· | ···················· | |
| M | 3894 hCoV-19/Wales/PHWC-47D8C6/2020\|EPI_ISL_586205\|2020 | ···················· | ···················· | ···················· | ···················· | ···················· | ···················· | |
| M | 3895 hCoV-19/Wales/PHWC-47D8D5/2020\|EPI_ISL_586206\|2020 | ···················· | ···················· | ···················· | ···················· | N··················· | ···················· | |
| M | 3896 hCoV-19/Wales/PHWC-47D8F3/2020\|EPI_ISL_586208\|2020 | ···················· | ·A·······A·········· | ···················· | ···················· | ···················· | ···················· | |
| M | 3897 hCoV-19/Wales/PHWC-47D91E/2020\|EPI_ISL_586209\|2020 | ···················· | ···················· | ···················· | ···················· | ···················· | ···················· | |
| M | 3899 hCoV-19/Wales/PHWC-47D94B/2020\|EPI_ISL_586212\|2020 | ···················· | ····A··············· | ···················· | ···················· | ···················· | ···················· | |
| M | 3901 hCoV-19/England/QEUH-A1E1C6/2020\|EPI_ISL_587479\|20 | ···················· | ···················· | ···················· | ···················· | ···················· | ···················· | |
| M | 3902 hCoV-19/England/QEUH-A1E73D/2020\|EPI_ISL_587480\|20 | ···················· | ···················· | ···················· | ···················· | ···················· | ···················· | |
| M | 3903 hCoV-19/England/QEUH-A1DF45/2020\|EPI_ISL_587481\|20 | ···················· | ···················· | ···················· | ···················· | ···················· | ···················· | |
| M | 3904 hCoV-19/England/QEUH-A1E22D/2020\|EPI_ISL_587483\|20 | ···················· | ···················· | ···················· | ···················· | ···················· | ···················· | |
| M | 3905 hCoV-19/England/QEUH-A1E14E/2020\|EPI_ISL_587484\|20 | ···················· | ···················· | ···················· | ···················· | ···················· | ···················· | |
| M | 3906 hCoV-19/England/QEUH-A1EDA4/2020\|EPI_ISL_587485\|20 | ···················· | ···················· | ···················· | ···················· | ···················· | ···················· | |
| M | 3907 hCoV-19/England/QEUH-A21694/2020\|EPI_ISL_587487\|20 | ···················· | ···················· | ···················· | ···················· | ···················· | ···················· | |
| M | 3908 hCoV-19/England/QEUH-A1E75B/2020\|EPI_ISL_587488\|20 | ···················· | ···················· | ···················· | ···················· | ···················· | ···················· | |
| M | 3909 hCoV-19/England/QEUH-A1DFFA/2020\|EPI_ISL_587489\|20 | ···················· | ···················· | ···················· | ···················· | ···················· | ···················· | |
| M | 3910 hCoV-19/England/QEUH-A1EBD7/2020\|EPI_ISL_587490\|20 | ···················· | ···················· | ···················· | ···················· | ···················· | ···················· | |
| M | 3911 hCoV-19/England/QEUH-A1DC11/2020\|EPI_ISL_587491\|20 | ···················· | ···················· | ···················· | ···················· | ···················· | ···················· | |
| M | 3912 hCoV-19/England/QEUH-A1ED95/2020\|EPI_ISL_587492\|20 | ···················· | ···················· | ···················· | ···················· | ···················· | ···················· | |
| M | 3913 hCoV-19/England/QEUH-A1E5AC/2020\|EPI_ISL_587493\|20 | ···················· | ···················· | ···················· | ···················· | ···················· | ···················· | |
| M | 3914 hCoV-19/England/QEUH-A1DB50/2020\|EPI_ISL_587494\|20 | ···················· | ···················· | ···················· | ···················· | ···················· | ···················· | |
| M | 3915 hCoV-19/England/QEUH-A1E23C/2020\|EPI_ISL_587496\|20 | ···················· | ···················· | ···················· | ···················· | ···················· | ···················· | |
| M | 3916 hCoV-19/England/QEUH-A1E9CE/2020\|EPI_ISL_587497\|20 | ···················· | ···················· | ···················· | ···················· | ···················· | ···················· | |
| M | 3917 hCoV-19/England/QEUH-A1DCC6/2020\|EPI_ISL_587535\|20 | ···················· | ···················· | ···················· | ···················· | ···················· | ···················· | |
| M | 3918 hCoV-19/England/QEUH-A1E2E1/2020\|EPI_ISL_587536\|20 | ···················· | ···················· | ···················· | ···················· | ···················· | ···················· | |
| M | 3919 hCoV-19/England/QEUH-A21FA8/2020\|EPI_ISL_587538\|20 | ···················· | ···················· | ···················· | ···················· | ···················· | ···················· | |
| M | 3920 hCoV-19/England/QEUH-A21FC6/2020\|EPI_ISL_587539\|20 | ···················· | ···················· | ···················· | ···················· | ···················· | ···················· | |
| M | 3921 hCoV-19/England/QEUH-A223AB/2020\|EPI_ISL_587540\|20 | ···················· | ···················· | ···················· | ···················· | ···················· | ···················· | |
| M | 3922 hCoV-19/England/QEUH-A1DFDC/2020\|EPI_ISL_587541\|20 | ···················· | ···················· | ···················· | ···················· | ···················· | ···················· | |
| M | 3923 hCoV-19/England/QEUH-A1F15C/2020\|EPI_ISL_587542\|20 | ···················· | ···················· | ···················· | ···················· | ···················· | ···················· | |
| M | 3924 hCoV-19/England/QEUH-A1ED77/2020\|EPI_ISL_587543\|20 | ···················· | ···················· | ···················· | ···················· | ···················· | ···················· | |
| M | 3925 hCoV-19/England/QEUH-9F804E/2020\|EPI_ISL_588865\|20 | ···················· | ···················· | ···················· | ···················· | ···················· | ···················· | |
| M | 3926 hCoV-19/England/QEUH-9F56B8/2020\|EPI_ISL_588866\|20 | ···················· | ···················· | ···················· | ···················· | ···················· | ···················· | |
| M | 3927 hCoV-19/England/QEUH-9F59A0/2020\|EPI_ISL_588868\|20 | ···················· | ·A·················· | ···················· | ···················· | ···················· | ···················· | |
| M | 3928 hCoV-19/England/QEUH-9F7F7F/2020\|EPI_ISL_588869\|20 | ···················· | ···················· | ···················· | ···················· | ···················· | ···················· | |
| M | 3929 hCoV-19/England/QEUH-9F9335/2020\|EPI_ISL_588870\|20 | ···················· | ···················· | ···················· | ···················· | ···················· | ···················· | |
| M | 3930 hCoV-19/England/QEUH-9F8CFF/2020\|EPI_ISL_588871\|20 | ···················· | ···················· | ···················· | ···················· | ···················· | ···················· | |
| M | 3931 hCoV-19/England/QEUH-9F8BB6/2020\|EPI_ISL_588872\|20 | ···················· | ···················· | ···················· | ···················· | ···················· | ···················· | |
| M | 3932 hCoV-19/England/QEUH-9F8891/2020\|EPI_ISL_588873\|20 | ···················· | ···················· | ···················· | ···················· | ···················· | ···················· | |
| M | 3933 hCoV-19/England/QEUH-9F8354/2020\|EPI_ISL_588874\|20 | ···················· | ···················· | ···················· | ···················· | ···················· | ···················· | |
| M | 3934 hCoV-19/England/QEUH-9F5630/2020\|EPI_ISL_588875\|20 | ···················· | ···················· | ···················· | ···················· | ···················· | ···················· | |
| M | 3935 hCoV-19/England/QEUH-9F8336/2020\|EPI_ISL_588878\|20 | ···················· | ···················· | ···················· | ···················· | ···················· | ···················· | |
| M | 3936 hCoV-19/Australia/VIC12472/2020\|EPI_ISL_592000\|202 | ···················· | ···················· | ···················· | ···················· | ···················· | ···················· | |
| M | 3937 hCoV-19/Australia/VIC12474/2020\|EPI_ISL_592001\|202 | ···················· | ······A············· | ···················· | ···················· | ···················· | ···················· | |
| M | 3938 hCoV-19/Australia/VIC12478/2020\|EPI_ISL_592002\|202 | ···················· | ···················· | ···················· | ···················· | ···················· | ···················· | |
| M | 3939 hCoV-19/Australia/VIC12479/2020\|EPI_ISL_592003\|202 | ···················· | ···················· | ···················· | ···················· | ···················· | ···················· | |
| M | 3940 hCoV-19/Australia/VIC12480/2020\|EPI_ISL_592004\|202 | ···················· | ······A············· | ···················· | ···················· | ···················· | ···················· | |
| M | 3941 hCoV-19/Australia/VIC12481/2020\|EPI_ISL_592005\|202 | ···················· | ···················· | ···················· | ···················· | ···················· | ···················· | |
| M | 3942 hCoV-19/Australia/VIC12486/2020\|EPI_ISL_592006\|202 | ···················· | ···················· | ···················· | ···················· | ···················· | ···················· | |
| M | 3943 hCoV-19/Australia/VIC12487/2020\|EPI_ISL_592007\|202 | ···················· | ···················· | ···················· | ···················· | ···················· | ···················· | |
| M | 3944 hCoV-19/Australia/VIC12491/2020\|EPI_ISL_592008\|202 | ···················· | ···················· | ···················· | ···················· | ···················· | ···················· | |
| M | 3945 hCoV-19/Australia/VIC12492/2020\|EPI_ISL_592009\|202 | ···················· | ···················· | ···················· | ···················· | ···················· | ···················· | |
| M | 3946 hCoV-19/Australia/VIC12493/2020\|EPI_ISL_592010\|202 | ···················· | ···················· | ···················· | ···················· | ···················· | ···················· | |
| M | 3947 hCoV-19/Australia/VIC12496/2020\|EPI_ISL_592011\|202 | ···················· | ···················· | ···················· | ···················· | ···················· | ···················· | |
| M | 3948 hCoV-19/Australia/VIC12498/2020\|EPI_ISL_592012\|202 | ···················· | ···················· | ···················· | ···················· | ···················· | ···················· | |
| M | 3949 hCoV-19/Australia/VIC12517/2020\|EPI_ISL_592013\|202 | ···················· | ···················· | ···················· | ···················· | ···················· | ···················· | |
| M | 3950 hCoV-19/Australia/VIC12519/2020\|EPI_ISL_592014\|202 | ···················· | ···················· | ···················· | ···················· | ···················· | ···················· | |
| M | 3952 hCoV-19/Australia/VIC12528/2020\|EPI_ISL_592016\|202 | ···················· | ···················· | ···················· | ···················· | ···················· | ···················· | |
| M | 3953 hCoV-19/Australia/VIC12529/2020\|EPI_ISL_592017\|202 | ···················· | ···················· | ···················· | ···················· | ···················· | ···················· | |
| M | 3954 hCoV-19/Australia/VIC12532/2020\|EPI_ISL_592018\|202 | ···················· | ···················· | ···················· | ···················· | ···················· | ···················· | |
| M | 3955 hCoV-19/Australia/VIC12534/2020\|EPI_ISL_592019\|202 | ···················· | ···················· | ···················· | ···················· | ···················· | ···················· | |
| M | 3957 hCoV-19/Australia/VIC12542/2020\|EPI_ISL_592021\|202 | ···················· | ···················· | ···················· | ···················· | ···················· | ···················· | |
| M | 3959 hCoV-19/Australia/VIC15452/2020\|EPI_ISL_593310\|202 | ···················· | ···················· | ···················· | ···················· | ···················· | ···················· | |
| M | 3960 hCoV-19/Australia/VIC15456/2020\|EPI_ISL_593311\|202 | ···················· | ···················· | ···················· | ···················· | ···················· | ···················· | |
| M | 3961 hCoV-19/Australia/VIC15457/2020\|EPI_ISL_593312\|202 | ···················· | ···················· | ···················· | ···················· | ···················· | ···················· | |
| M | 3962 hCoV-19/Australia/VIC15458/2020\|EPI_ISL_593313\|202 | ···················· | ···················· | ···················· | ···················· | ···················· | ···················· | |

| | | | | | | | | |
|---|---|---|---|---|---|---|---|---|
| M | 3963 hCoV-19/Australia/VIC15459/2020\|EPI_ISL_593314\|202 | ···················· | ···················· | ···················· | ···················· | ··················NN | ···················· | 1 |
| M | 3964 hCoV-19/Australia/VIC15460/2020\|EPI_ISL_593315\|202 | ···················· | ···················· | ···················· | ···················· | ···················· | ···················· | |
| M | 3965 hCoV-19/Australia/VIC15462/2020\|EPI_ISL_593316\|202 | ···················· | ···················· | ···················· | ···················· | ···················· | ···················· | |
| M | 3966 hCoV-19/Australia/VIC15463/2020\|EPI_ISL_593317\|202 | ···················· | ···················· | ···················· | ···················· | ···················· | ···················· | |
| M | 3967 hCoV-19/Australia/VIC15465/2020\|EPI_ISL_593318\|202 | ···················· | ···················· | ···················· | ···················· | ···················· | ···················· | |
| M | 3968 hCoV-19/Australia/VIC15468/2020\|EPI_ISL_593319\|202 | ···················· | ···················· | ···················· | ···················· | ···················· | ···················· | |
| M | 3969 hCoV-19/Australia/VIC15469/2020\|EPI_ISL_593320\|202 | ···················· | ···················· | ···················· | ···················· | ···················· | ···················· | |
| M | 3970 hCoV-19/Australia/VIC15472/2020\|EPI_ISL_593321\|202 | ···················· | ···················· | ···················· | ···················· | ···················· | ···················· | |
| M | 3971 hCoV-19/Australia/VIC15475/2020\|EPI_ISL_593322\|202 | ···················· | ···················· | ···················· | ···················· | ···················· | ···················· | |
| M | 3972 hCoV-19/Australia/VIC15476/2020\|EPI_ISL_593323\|202 | ···················· | ···················· | ···················· | ···················· | ···················· | ···················· | |
| M | 3973 hCoV-19/Australia/VIC15479/2020\|EPI_ISL_593324\|202 | ···················· | ···················· | ···················· | ···················· | ···················· | ···················· | |
| M | 3974 hCoV-19/Australia/VIC15482/2020\|EPI_ISL_593325\|202 | ···················· | ···················· | ···················· | ···················· | ···················· | ···················· | |
| M | 3975 hCoV-19/Australia/VIC15483/2020\|EPI_ISL_593326\|202 | ···················· | ···················· | ···················· | ···················· | ···················· | ···················· | |
| M | 3976 hCoV-19/Australia/VIC15484/2020\|EPI_ISL_593327\|202 | ···················· | ···················· | ···················· | ···················· | ···················· | ···················· | |
| M | 3977 hCoV-19/Australia/VIC15488/2020\|EPI_ISL_593328\|202 | ···················· | ···················· | ···················· | ···················· | ···················· | ···················· | |
| M | 3978 hCoV-19/Australia/VIC15489/2020\|EPI_ISL_593329\|202 | ···················· | ···················· | ···················· | ···················· | ···················· | ···················· | |
| M | 3979 hCoV-19/Australia/VIC15490/2020\|EPI_ISL_593330\|202 | ···················· | ···················· | ···················· | ···················· | ···················· | ···················· | |
| M | 3980 hCoV-19/Australia/VIC15491/2020\|EPI_ISL_593331\|202 | ···················· | ···················· | ···················· | ···················· | ···················· | ···················· | |
| M | 3981 hCoV-19/Australia/VIC15493/2020\|EPI_ISL_593332\|202 | ···················· | ···················· | ···················· | ···················· | ···················· | ···················· | |
| M | 3982 hCoV-19/Australia/VIC15494/2020\|EPI_ISL_593333\|202 | ···················· | ···················· | ···················· | ···················· | ···················· | ···················· | |
| M | 3983 hCoV-19/Australia/VIC15495/2020\|EPI_ISL_593334\|202 | ···················· | ···················· | ···················· | ···················· | ···················· | ···················· | |
| M | 3984 hCoV-19/Australia/VIC15497/2020\|EPI_ISL_593335\|202 | ···················· | ···················· | ···················· | ···················· | ···················· | ···················· | |
| M | 3985 hCoV-19/Australia/VIC15499/2020\|EPI_ISL_593336\|202 | ···················· | ···················· | ···················· | ···················· | ···················· | ···················· | |
| M | 3986 hCoV-19/Australia/VIC15500/2020\|EPI_ISL_593337\|202 | ···················· | ···················· | ···················· | ···················· | ···················· | ···················· | |
| M | 3987 hCoV-19/Australia/VIC15503/2020\|EPI_ISL_593338\|202 | ···················· | ···················· | ···················· | ···················· | ···················· | ···················· | |
| M | 3988 hCoV-19/Australia/VIC15505/2020\|EPI_ISL_593339\|202 | ···················· | ···················· | ···················· | ···················· | ···················· | ···················· | |
| M | 3989 hCoV-19/Australia/VIC15506/2020\|EPI_ISL_593340\|202 | ···················· | ···················· | ···················· | ···················· | ···················· | ···················· | |
| M | 3990 hCoV-19/Australia/VIC15507/2020\|EPI_ISL_593341\|202 | ···················· | ···················· | ···················· | ···················· | ···················· | ···················· | |
| M | 3992 hCoV-19/Australia/VIC15512/2020\|EPI_ISL_593343\|202 | ···················· | ···················· | ···················· | ···················· | ···················· | ···················· | |
| M | 3993 hCoV-19/Australia/VIC15513/2020\|EPI_ISL_593344\|202 | ···················· | ···················· | ···················· | ···················· | ···················· | ···················· | |
| M | 3994 hCoV-19/Australia/VIC15518/2020\|EPI_ISL_593345\|202 | ···················· | ···················· | ···················· | ···················· | ···················· | ···················· | |
| M | 3995 hCoV-19/Australia/VIC15519/2020\|EPI_ISL_593346\|202 | ···················· | ···················· | ···················· | ···················· | ···················· | ···················· | |
| M | 3996 hCoV-19/Australia/VIC15520/2020\|EPI_ISL_593347\|202 | ···················· | ···················· | ···················· | ···················· | ···················· | ···················· | |
| M | 3997 hCoV-19/Australia/VIC15523/2020\|EPI_ISL_593348\|202 | ···················· | ···················· | ···················· | ···················· | ···················· | ···················· | |
| M | 3998 hCoV-19/Australia/VIC15530/2020\|EPI_ISL_593349\|202 | ···················· | ···················· | ···················· | ···················· | ···················· | ···················· | |
| M | 3999 hCoV-19/Australia/VIC15534/2020\|EPI_ISL_593350\|202 | ···················· | ···················· | ···················· | ···················· | ···················· | ···················· | |
| M | 4000 hCoV-19/Australia/VIC15535/2020\|EPI_ISL_593351\|202 | ···················· | ···················· | ···················· | ···················· | N··················· | ···················· | |
| M | 4001 hCoV-19/Australia/VIC15536/2020\|EPI_ISL_593352\|202 | ···················· | ···················· | ···················· | ···················· | ···················· | ···················· | |
| M | 4003 hCoV-19/England/BIRM-65A38/2020\|EPI_ISL_594603\|202 | ···················· | ···················· | ···················· | ···················· | ···················· | ···················· | |
| M | 4006 hCoV-19/England/BIRM-65A65/2020\|EPI_ISL_594606\|202 | ···················· | ···················· | ···················· | ···················· | ···················· | ···················· | |
| M | 4007 hCoV-19/England/BIRM-65A83/2020\|EPI_ISL_594607\|202 | ···················· | ···················· | ···················· | ···················· | ···················· | ···················· | |
| M | 4008 hCoV-19/England/BIRM-65A92/2020\|EPI_ISL_594608\|202 | ···················· | ···················· | ···················· | ···················· | ···················· | ···················· | |
| M | 4009 hCoV-19/England/BIRM-65F39/2020\|EPI_ISL_594609\|202 | ···················· | ···················· | ···················· | ···················· | ···················· | ···················· | |
| M | 4016 hCoV-19/England/CAMC-9EC766/2020\|EPI_ISL_594617\|20 | ···················· | ···················· | ···················· | ···················· | ···················· | ···················· | |
| M | 4017 hCoV-19/England/CAMC-9EC7B1/2020\|EPI_ISL_594618\|20 | ···················· | ···················· | ···················· | ···················· | ···················· | ···················· | |
| M | 4018 hCoV-19/England/CAMC-9EC7C0/2020\|EPI_ISL_594619\|20 | ···················· | ···················· | ···················· | ···················· | ···················· | ···················· | |
| M | 4019 hCoV-19/England/CAMC-9ECAA9/2020\|EPI_ISL_594638\|20 | ···················· | ···················· | ···················· | ···················· | ···················· | ···················· | |
| M | 4020 hCoV-19/England/CAMC-9ECB1F/2020\|EPI_ISL_594640\|20 | ···················· | ···················· | ···················· | ···················· | ···················· | ···················· | |
| M | 4021 hCoV-19/England/CAMC-9ECB5B/2020\|EPI_ISL_594641\|20 | ···················· | ···················· | ···················· | ···················· | ···················· | ···················· | |
| M | 4022 hCoV-19/England/CAMC-9ECB79/2020\|EPI_ISL_594642\|20 | ···················· | ···················· | ···················· | ···················· | ···················· | ···················· | |
| M | 4023 hCoV-19/England/CAMC-9ECB97/2020\|EPI_ISL_594643\|20 | ···················· | ···················· | ···················· | ···················· | ···················· | ···················· | |
| M | 4024 hCoV-19/England/CAMC-9ECBA6/2020\|EPI_ISL_594644\|20 | ···················· | ···················· | ···················· | ···················· | ···················· | ···················· | |
| M | 4025 hCoV-19/Scotland/CVR4843/2020\|EPI_ISL_594646\|2020- | ···················· | ···················· | ···················· | ···················· | ···················· | ···················· | |
| M | 4027 hCoV-19/Scotland/CVR4854/2020\|EPI_ISL_594654\|2020- | ···················· | ···················· | ···················· | ···················· | ··············N····· | N··················· | |
| M | 4028 hCoV-19/Scotland/CVR4889/2020\|EPI_ISL_594656\|2020- | ···················· | ···················· | ···················· | ···················· | ···················· | ···················· | |
| M | 4029 hCoV-19/Scotland/CVR4895/2020\|EPI_ISL_594662\|2020- | ···················· | ···················· | ···················· | ···················· | ···················· | ···················· | |
| M | 4030 hCoV-19/Scotland/CVR4898/2020\|EPI_ISL_594665\|2020- | ···················· | ···················· | ···················· | ···················· | ···················· | ···················· | |
| M | 4031 hCoV-19/Scotland/CVR4899/2020\|EPI_ISL_594666\|2020- | ···················· | ···················· | ···················· | ···················· | ···················· | ···················· | |
| M | 4032 hCoV-19/Scotland/CVR4900/2020\|EPI_ISL_594667\|2020- | ···················· | ···················· | ···················· | ···················· | ···················· | NN·················· | |
| M | 4033 hCoV-19/Scotland/CVR4901/2020\|EPI_ISL_594668\|2020- | ···················· | ···················· | ···················· | ···················· | ···················· | ···················· | |
| M | 4034 hCoV-19/Scotland/CVR4903/2020\|EPI_ISL_594670\|2020- | ···················· | ···················· | ···················· | ···················· | ···················· | ···················· | |
| M | 4035 hCoV-19/Scotland/CVR4905/2020\|EPI_ISL_594672\|2020- | ···················· | ···················· | ···················· | ···················· | ···················· | ···················· | |
| M | 4036 hCoV-19/Scotland/CVR4907/2020\|EPI_ISL_594674\|2020- | ···················· | ···················· | ···················· | ···················· | ···················· | ···················· | |
| M | 4037 hCoV-19/Scotland/CVR4910/2020\|EPI_ISL_594676\|2020- | ···················· | ···················· | ···················· | ···················· | ···················· | ···················· | |
| M | 4038 hCoV-19/Scotland/CVR4911/2020\|EPI_ISL_594677\|2020- | ···················· | ···················· | ···················· | ···················· | ··············N····· | NN·················· | |
| M | 4039 hCoV-19/Scotland/CVR4913/2020\|EPI_ISL_594679\|2020- | ···················· | ···················· | ···················· | ···················· | ···················· | ···················· | |
| M | 4040 hCoV-19/Scotland/CVR4916/2020\|EPI_ISL_594682\|2020- | ···················· | ···················· | ···················· | ···················· | ···················· | ···················· | |
| M | 4044 hCoV-19/England/QEUH-A31585/2020\|EPI_ISL_595903\|20 | ···················· | ···················· | ···················· | ···················· | ···················· | ···················· | |
| M | 4045 hCoV-19/England/QEUH-A31594/2020\|EPI_ISL_595904\|20 | ···················· | ···················· | ···················· | ···················· | ···················· | ···················· | |
| M | 4046 hCoV-19/England/QEUH-A315A3/2020\|EPI_ISL_595905\|20 | ···················· | ···················· | ···················· | ···················· | ···················· | ···················· | |
| M | 4047 hCoV-19/England/QEUH-A31752/2020\|EPI_ISL_595906\|20 | ···················· | ···················· | ···················· | ···················· | ···················· | ···················· | |
| M | 4049 hCoV-19/England/QEUH-A3179E/2020\|EPI_ISL_595909\|20 | ···················· | ···················· | ···················· | ···················· | ···················· | ···················· | |
| M | 4050 hCoV-19/England/QEUH-A317AD/2020\|EPI_ISL_595910\|20 | ···················· | ···················· | ···················· | ···················· | ···················· | ···················· | |
| M | 4051 hCoV-19/England/QEUH-A317BC/2020\|EPI_ISL_595911\|20 | ···················· | ···················· | ···················· | ···················· | ···················· | ···················· | |
| M | 4052 hCoV-19/England/QEUH-A317CB/2020\|EPI_ISL_595912\|20 | ···················· | ···················· | ···················· | ···················· | ···················· | ···················· | |
| M | 4054 hCoV-19/Scotland/QEUH-A31A77/2020\|EPI_ISL_595914\|2 | ···················· | ···················· | ···················· | ···················· | ···················· | ···················· | |
| M | 4055 hCoV-19/England/QEUH-A48CB3/2020\|EPI_ISL_595916\|20 | ···················· | ···················· | ···················· | ···················· | ······T············· | ···················· | |
| M | 4060 hCoV-19/Scotland/QEUH-A48E08/2020\|EPI_ISL_595926\|2 | ···················· | ···················· | ···················· | ···················· | ···················· | ···················· | |
| M | 4061 hCoV-19/Scotland/QEUH-A48E17/2020\|EPI_ISL_595927\|2 | ···················· | ···················· | ···················· | ···················· | ···················· | ···················· | |
| M | 4062 hCoV-19/Scotland/QEUH-A48E44/2020\|EPI_ISL_595929\|2 | ···················· | ···················· | ···················· | ···················· | ···················· | ···················· | |

<table>
<tr><td>M</td><td>4063</td><td>hCoV-19/Scotland/QEUH-A48E53/2020|EPI_ISL_595930|2</td><td>····················</td><td>····················</td><td>·····················</td><td>····················</td><td>····················</td><td>····················</td><td></td></tr>
<tr><td>M</td><td>4064</td><td>hCoV-19/Scotland/QEUH-A48E62/2020|EPI_ISL_595931|2</td><td>····················</td><td>····················</td><td>·····················</td><td>····················</td><td>····················</td><td>····················</td><td></td></tr>
<tr><td>M</td><td>4065</td><td>hCoV-19/Scotland/QEUH-A48EAE/2020|EPI_ISL_595935|2</td><td>····················</td><td>····················</td><td>·····················</td><td>····················</td><td>····················</td><td>····················</td><td></td></tr>
<tr><td>M</td><td>4066</td><td>hCoV-19/Scotland/QEUH-A48EBD/2020|EPI_ISL_595936|2</td><td>····················</td><td>····················</td><td>·····················</td><td>····················</td><td>····················</td><td>····················</td><td></td></tr>
<tr><td>M</td><td>4067</td><td>hCoV-19/Scotland/QEUH-A48EDB/2020|EPI_ISL_595937|2</td><td>····················</td><td>····················</td><td>·····················</td><td>····················</td><td>····················</td><td>····················</td><td></td></tr>
<tr><td>M</td><td>4068</td><td>hCoV-19/England/QEUH-A48EEA/2020|EPI_ISL_595938|20</td><td>····················</td><td>····················</td><td>·····················</td><td>····················</td><td>····················</td><td>····················</td><td></td></tr>
<tr><td>M</td><td>4070</td><td>hCoV-19/Scotland/QEUH-A48FAB/2020|EPI_ISL_595941|2</td><td>····················</td><td>····················</td><td>·····················</td><td>····················</td><td>····················</td><td>····················</td><td></td></tr>
<tr><td>M</td><td>4071</td><td>hCoV-19/Scotland/QEUH-A48FBA/2020|EPI_ISL_595942|2</td><td>····················</td><td>····················</td><td>·····················</td><td>····················</td><td>····················</td><td>····················</td><td></td></tr>
<tr><td>M</td><td>4072</td><td>hCoV-19/Scotland/QEUH-A48FD8/2020|EPI_ISL_595943|2</td><td>····················</td><td>····················</td><td>·····················</td><td>····················</td><td>····················</td><td>····················</td><td></td></tr>
<tr><td>M</td><td>4073</td><td>hCoV-19/Scotland/QEUH-A49001/2020|EPI_ISL_595945|2</td><td>····················</td><td>····················</td><td>·····················</td><td>····················</td><td>····················</td><td>····················</td><td></td></tr>
<tr><td>M</td><td>4074</td><td>hCoV-19/Scotland/QEUH-A4902F/2020|EPI_ISL_595947|2</td><td>····················</td><td>····················</td><td>·····················</td><td>····················</td><td>····················</td><td>····················</td><td></td></tr>
<tr><td>M</td><td>4075</td><td>hCoV-19/Scotland/QEUH-A4903E/2020|EPI_ISL_595948|2</td><td>····················</td><td>····················</td><td>·····················</td><td>····················</td><td>····················</td><td>····················</td><td></td></tr>
<tr><td>M</td><td>4076</td><td>hCoV-19/Scotland/QEUH-A4AA8A/2020|EPI_ISL_595968|2</td><td>····················</td><td>····················</td><td>·····················</td><td>····················</td><td>····················</td><td>····················</td><td></td></tr>
<tr><td>M</td><td>4077</td><td>hCoV-19/Scotland/QEUH-A4AAD5/2020|EPI_ISL_595971|2</td><td>····················</td><td>····················</td><td>·····················</td><td>····················</td><td>····················</td><td>····················</td><td></td></tr>
<tr><td>M</td><td>4078</td><td>hCoV-19/Scotland/QEUH-A4C52D/2020|EPI_ISL_595974|2</td><td>····················</td><td>····················</td><td>·····················</td><td>····················</td><td>····················</td><td>····················</td><td></td></tr>
<tr><td>M</td><td>4079</td><td>hCoV-19/Scotland/QEUH-A4C55A/2020|EPI_ISL_595976|2</td><td>····················</td><td>····················</td><td>·····················</td><td>····················</td><td>····················</td><td>····················</td><td></td></tr>
<tr><td>M</td><td>4080</td><td>hCoV-19/Scotland/QEUH-A4C569/2020|EPI_ISL_595977|2</td><td>····················</td><td>····················</td><td>·····················</td><td>····················</td><td>····················</td><td>····················</td><td></td></tr>
<tr><td>M</td><td>4081</td><td>hCoV-19/England/QEUH-A4C578/2020|EPI_ISL_595978|20</td><td>····················</td><td>····················</td><td>·····················</td><td>····················</td><td>····················</td><td>····················</td><td></td></tr>
<tr><td>M</td><td>4085</td><td>hCoV-19/England/QEUH-A4C5B4/2020|EPI_ISL_595982|20</td><td>····················</td><td>····················</td><td>·····················</td><td>····················</td><td>····················</td><td>····················</td><td></td></tr>
<tr><td>M</td><td>4088</td><td>hCoV-19/Scotland/QEUH-A4C693/2020|EPI_ISL_595988|2</td><td>····················</td><td>····················</td><td>·····················</td><td>····················</td><td>····················</td><td>····················</td><td></td></tr>
<tr><td>M</td><td>4089</td><td>hCoV-19/England/QEUH-A4C6A2/2020|EPI_ISL_595989|20</td><td>····················</td><td>····················</td><td>·····················</td><td>····················</td><td>NNN·················</td><td>····················</td><td>1</td></tr>
<tr><td>M</td><td>4090</td><td>hCoV-19/Scotland/QEUH-A4C6C0/2020|EPI_ISL_595990|2</td><td>····················</td><td>····················</td><td>·····················</td><td>····················</td><td>······T·············</td><td>····················</td><td></td></tr>
<tr><td>M</td><td>4092</td><td>hCoV-19/Scotland/QEUH-A4C709/2020|EPI_ISL_595993|2</td><td>····················</td><td>····················</td><td>·····················</td><td>····················</td><td>····················</td><td>····················</td><td></td></tr>
<tr><td>M</td><td>4093</td><td>hCoV-19/England/QEUH-A4C727/2020|EPI_ISL_595995|20</td><td>····················</td><td>····················</td><td>·····················</td><td>····················</td><td>····················</td><td>····················</td><td></td></tr>
<tr><td>M</td><td>4094</td><td>hCoV-19/Scotland/QEUH-A4C745/2020|EPI_ISL_595997|2</td><td>····················</td><td>····················</td><td>·····················</td><td>····················</td><td>····················</td><td>····················</td><td></td></tr>
<tr><td>M</td><td>4095</td><td>hCoV-19/Scotland/QEUH-A4C754/2020|EPI_ISL_595998|2</td><td>····················</td><td>····················</td><td>·····················</td><td>····················</td><td>····················</td><td>····················</td><td></td></tr>
<tr><td>M</td><td>4097</td><td>hCoV-19/Wales/PHWC-47D86C/2020|EPI_ISL_586201|2020</td><td>····················</td><td>····················</td><td>·····················</td><td>····················</td><td>····················</td><td>····················</td><td></td></tr>
<tr><td>M</td><td>4100</td><td>hCoV-19/Wales/PHWC-47D978/2020|EPI_ISL_586215|2020</td><td>····················</td><td>····················</td><td>·····················</td><td>····················</td><td>····················</td><td>····················</td><td></td></tr>
<tr><td>M</td><td>4101</td><td>hCoV-19/Wales/PHWC-47D987/2020|EPI_ISL_586216|2020</td><td>····················</td><td>····················</td><td>·····················</td><td>····················</td><td>····················</td><td>····················</td><td></td></tr>
<tr><td>M</td><td>4102</td><td>hCoV-19/Wales/PHWC-47D9A5/2020|EPI_ISL_586218|2020</td><td>····················</td><td>····················</td><td>·····················</td><td>····················</td><td>····················</td><td>····················</td><td></td></tr>
<tr><td>M</td><td>4103</td><td>hCoV-19/Wales/PHWC-47D9B4/2020|EPI_ISL_586219|2020</td><td>····················</td><td>····················</td><td>·····················</td><td>····················</td><td>····················</td><td>····················</td><td></td></tr>
<tr><td>M</td><td>4105</td><td>hCoV-19/Wales/PHWC-47D9E1/2020|EPI_ISL_586222|2020</td><td>····················</td><td>····················</td><td>·····················</td><td>····················</td><td>····················</td><td>····················</td><td></td></tr>
<tr><td>M</td><td>4106</td><td>hCoV-19/Wales/PHWC-47D9F0/2020|EPI_ISL_586223|2020</td><td>····················</td><td>····················</td><td>·····················</td><td>····················</td><td>NNN·················</td><td>····················</td><td>1</td></tr>
<tr><td>M</td><td>4108</td><td>hCoV-19/Wales/PHWC-47DA2A/2020|EPI_ISL_586226|2020</td><td>····················</td><td>····················</td><td>·····················</td><td>····················</td><td>····················</td><td>····················</td><td></td></tr>
<tr><td>M</td><td>4109</td><td>hCoV-19/Wales/PHWC-47DA48/2020|EPI_ISL_586228|2020</td><td>····················</td><td>····················</td><td>·····················</td><td>····················</td><td>····················</td><td>····················</td><td></td></tr>
<tr><td>M</td><td>4111</td><td>hCoV-19/Wales/PHWC-47DA93/2020|EPI_ISL_586233|2020</td><td>····················</td><td>····················</td><td>·····················</td><td>····················</td><td>····················</td><td>····················</td><td></td></tr>
<tr><td>M</td><td>4113</td><td>hCoV-19/Wales/PHWC-47DAB1/2020|EPI_ISL_586235|2020</td><td>····················</td><td>····················</td><td>·····················</td><td>····················</td><td>····················</td><td>····················</td><td></td></tr>
<tr><td>M</td><td>4116</td><td>hCoV-19/Wales/PHWC-47DAFD/2020|EPI_ISL_586239|2020</td><td>····················</td><td>····················</td><td>·····················</td><td>····················</td><td>····················</td><td>····················</td><td></td></tr>
<tr><td>M</td><td>4117</td><td>hCoV-19/USA/AK-PHL573/2020|EPI_ISL_586243|2020-06-</td><td>····················</td><td>····················</td><td>·····················</td><td>····················</td><td>····················</td><td>····················</td><td></td></tr>
<tr><td>M</td><td>4118</td><td>hCoV-19/USA/AK-PHL592/2020|EPI_ISL_586244|2020-06-</td><td>····················</td><td>····················</td><td>·····················</td><td>····················</td><td>····················</td><td>····················</td><td></td></tr>
<tr><td>M</td><td>4119</td><td>hCoV-19/USA/AK-PHL602/2020|EPI_ISL_586245|2020-06-</td><td>····················</td><td>·A··················</td><td>·····················</td><td>····················</td><td>····················</td><td>····················</td><td></td></tr>
<tr><td>M</td><td>4120</td><td>hCoV-19/USA/AK-PHL627/2020|EPI_ISL_586246|2020-06-</td><td>····················</td><td>····················</td><td>·····················</td><td>····················</td><td>····················</td><td>····················</td><td></td></tr>
<tr><td>M</td><td>4121</td><td>hCoV-19/USA/AK-PHL635/2020|EPI_ISL_586247|2020-06-</td><td>····················</td><td>····················</td><td>·····················</td><td>····················</td><td>····················</td><td>····················</td><td></td></tr>
<tr><td>M</td><td>4122</td><td>hCoV-19/USA/AK-PHL640/2020|EPI_ISL_586248|2020-06-</td><td>····················</td><td>····················</td><td>·····················</td><td>····················</td><td>····················</td><td>····················</td><td></td></tr>
<tr><td>M</td><td>4123</td><td>hCoV-19/USA/AK-PHL646/2020|EPI_ISL_586249|2020-06-</td><td>····················</td><td>····················</td><td>·····················</td><td>····················</td><td>····················</td><td>····················</td><td></td></tr>
<tr><td>M</td><td>4124</td><td>hCoV-19/USA/AK-PHL647/2020|EPI_ISL_586250|2020-06-</td><td>····················</td><td>····················</td><td>·····················</td><td>····················</td><td>····················</td><td>····················</td><td></td></tr>
<tr><td>M</td><td>4125</td><td>hCoV-19/USA/AK-PHL648/2020|EPI_ISL_586251|2020-06-</td><td>····················</td><td>····················</td><td>·····················</td><td>····················</td><td>····················</td><td>····················</td><td></td></tr>
<tr><td>M</td><td>4126</td><td>hCoV-19/USA/AK-PHL660/2020|EPI_ISL_586252|2020-06-</td><td>····················</td><td>····················</td><td>·····················</td><td>····················</td><td>····················</td><td>····················</td><td></td></tr>
<tr><td>M</td><td>4127</td><td>hCoV-19/USA/AK-PHL672/2020|EPI_ISL_586253|2020-06-</td><td>····················</td><td>····················</td><td>·····················</td><td>····················</td><td>····················</td><td>····················</td><td></td></tr>
<tr><td>M</td><td>4128</td><td>hCoV-19/USA/AK-PHL676/2020|EPI_ISL_586254|2020-06-</td><td>····················</td><td>····················</td><td>·····················</td><td>····················</td><td>····················</td><td>····················</td><td></td></tr>
<tr><td>M</td><td>4129</td><td>hCoV-19/USA/AK-PHL683/2020|EPI_ISL_586255|2020-06-</td><td>····················</td><td>····················</td><td>·····················</td><td>····················</td><td>····················</td><td>····················</td><td></td></tr>
<tr><td>M</td><td>4130</td><td>hCoV-19/USA/AK-PHL689/2020|EPI_ISL_586256|2020-06-</td><td>····················</td><td>····················</td><td>·····················</td><td>····················</td><td>····················</td><td>····················</td><td></td></tr>
<tr><td>M</td><td>4131</td><td>hCoV-19/USA/AK-PHL699/2020|EPI_ISL_586257|2020-06-</td><td>····················</td><td>····················</td><td>·····················</td><td>····················</td><td>····················</td><td>····················</td><td></td></tr>
<tr><td>M</td><td>4132</td><td>hCoV-19/USA/AK-PHL706/2020|EPI_ISL_586258|2020-06-</td><td>····················</td><td>····················</td><td>·····················</td><td>····················</td><td>····················</td><td>····················</td><td></td></tr>
<tr><td>M</td><td>4133</td><td>hCoV-19/USA/AK-PHL707/2020|EPI_ISL_586259|2020-06-</td><td>····················</td><td>····················</td><td>·····················</td><td>····················</td><td>····················</td><td>····················</td><td></td></tr>
<tr><td>M</td><td>4134</td><td>hCoV-19/USA/AK-PHL727/2020|EPI_ISL_586260|2020-06-</td><td>····················</td><td>·A··················</td><td>·····················</td><td>····················</td><td>····················</td><td>····················</td><td></td></tr>
<tr><td>M</td><td>4135</td><td>hCoV-19/USA/AK-PHL734/2020|EPI_ISL_586261|2020-06-</td><td>····················</td><td>····················</td><td>·····················</td><td>····················</td><td>····················</td><td>····················</td><td></td></tr>
<tr><td>M</td><td>4136</td><td>hCoV-19/USA/AK-PHL737/2020|EPI_ISL_586262|2020-06-</td><td>····················</td><td>····················</td><td>·····················</td><td>····················</td><td>····················</td><td>····················</td><td></td></tr>
<tr><td>M</td><td>4137</td><td>hCoV-19/USA/AK-PHL760/2020|EPI_ISL_586263|2020-07-</td><td>····················</td><td>····················</td><td>·····················</td><td>····················</td><td>····················</td><td>····················</td><td></td></tr>
<tr><td>M</td><td>4138</td><td>hCoV-19/USA/AK-PHL767/2020|EPI_ISL_586264|2020-07-</td><td>····················</td><td>····················</td><td>·····················</td><td>····················</td><td>····················</td><td>····················</td><td></td></tr>
<tr><td>M</td><td>4139</td><td>hCoV-19/USA/AK-PHL796/2020|EPI_ISL_586265|2020-07-</td><td>····················</td><td>····················</td><td>·····················</td><td>····················</td><td>····················</td><td>····················</td><td></td></tr>
<tr><td>M</td><td>4140</td><td>hCoV-19/USA/AK-PHL806/2020|EPI_ISL_586266|2020-07-</td><td>····················</td><td>····················</td><td>·····················</td><td>····················</td><td>····················</td><td>····················</td><td></td></tr>
<tr><td>M</td><td>4141</td><td>hCoV-19/Canada/ON-CVD00060/2020|EPI_ISL_586268|202</td><td>····················</td><td>····················</td><td>·····················</td><td>····················</td><td>····················</td><td>····················</td><td></td></tr>
<tr><td>M</td><td>4142</td><td>hCoV-19/England/QEUH-A1DF72/2020|EPI_ISL_587512|20</td><td>····················</td><td>····················</td><td>·····················</td><td>····················</td><td>····T···············</td><td>····················</td><td></td></tr>
<tr><td>M</td><td>4143</td><td>hCoV-19/England/QEUH-A1DC20/2020|EPI_ISL_587513|20</td><td>····················</td><td>····················</td><td>·····················</td><td>····················</td><td>····················</td><td>····················</td><td></td></tr>
<tr><td>M</td><td>4144</td><td>hCoV-19/England/QEUH-A21F4E/2020|EPI_ISL_587514|20</td><td>····················</td><td>····················</td><td>·····················</td><td>····················</td><td>····················</td><td>····················</td><td></td></tr>
<tr><td>M</td><td>4145</td><td>hCoV-19/England/QEUH-A213CA/2020|EPI_ISL_587516|20</td><td>····················</td><td>····················</td><td>·····················</td><td>····················</td><td>····················</td><td>····················</td><td></td></tr>
<tr><td>M</td><td>4146</td><td>hCoV-19/England/QEUH-A21306/2020|EPI_ISL_587517|20</td><td>····················</td><td>····················</td><td>·····················</td><td>····················</td><td>····················</td><td>····················</td><td></td></tr>
<tr><td>M</td><td>4147</td><td>hCoV-19/England/QEUH-A1E4DC/2020|EPI_ISL_587518|20</td><td>····················</td><td>····················</td><td>·····················</td><td>····················</td><td>····················</td><td>····················</td><td></td></tr>
<tr><td>M</td><td>4148</td><td>hCoV-19/England/QEUH-A22305/2020|EPI_ISL_587519|20</td><td>····················</td><td>····················</td><td>·····················</td><td>····················</td><td>····················</td><td>····················</td><td></td></tr>
<tr><td>M</td><td>4149</td><td>hCoV-19/England/QEUH-A1DDC3/2020|EPI_ISL_587520|20</td><td>····················</td><td>····················</td><td>·····················</td><td>····················</td><td>····················</td><td>····················</td><td></td></tr>
<tr><td>M</td><td>4150</td><td>hCoV-19/England/QEUH-A1F1A7/2020|EPI_ISL_587521|20</td><td>····················</td><td>····················</td><td>·····················</td><td>····················</td><td>····················</td><td>····················</td><td></td></tr>
<tr><td>M</td><td>4151</td><td>hCoV-19/England/QEUH-A1E16C/2020|EPI_ISL_587522|20</td><td>····················</td><td>·A··················</td><td>·····················</td><td>····················</td><td>····················</td><td>····················</td><td></td></tr>
<tr><td>M</td><td>4152</td><td>hCoV-19/England/QEUH-A1F17A/2020|EPI_ISL_587523|20</td><td>····················</td><td>····················</td><td>·····················</td><td>····················</td><td>····················</td><td>····················</td><td></td></tr>
<tr><td>M</td><td>4153</td><td>hCoV-19/England/QEUH-A1DB05/2020|EPI_ISL_587524|20</td><td>····················</td><td>····················</td><td>·····················</td><td>····················</td><td>····················</td><td>····················</td><td></td></tr>
<tr><td>M</td><td>4154</td><td>hCoV-19/England/QEUH-A1F480/2020|EPI_ISL_587526|20</td><td>····················</td><td>····················</td><td>·····················</td><td>····················</td><td>····················</td><td>····················</td><td></td></tr>
<tr><td>M</td><td>4155</td><td>hCoV-19/England/QEUH-A1E2D2/2020|EPI_ISL_587527|20</td><td>····················</td><td>····················</td><td>·····················</td><td>····················</td><td>····················</td><td>····················</td><td></td></tr>
<tr><td>M</td><td>4156</td><td>hCoV-19/England/QEUH-A1DE93/2020|EPI_ISL_587528|20</td><td>····················</td><td>····················</td><td>·····················</td><td>····················</td><td>····················</td><td>····················</td><td></td></tr>
<tr><td>M</td><td>4157</td><td>hCoV-19/England/QEUH-A223BA/2020|EPI_ISL_587529|20</td><td>····················</td><td>····················</td><td>·····················</td><td>····················</td><td>····················</td><td>····················</td><td></td></tr>
<tr><td>M</td><td>4158</td><td>hCoV-19/England/QEUH-A21175/2020|EPI_ISL_587530|20</td><td>····················</td><td>····················</td><td>·····················</td><td>····················</td><td>····················</td><td>····················</td><td></td></tr>
</table>

| | | | | | | | | |
|---|---|---|---|---|---|---|---|---|
| M | 4159 | hCoV-19/England/QEUH-A1EFCC/2020\|EPI_ISL_587532\|20 | ···················· | ···················· | ···················· | ···················· | ···················· | ···················· |
| M | 4160 | hCoV-19/England/QEUH-A1E8EF/2020\|EPI_ISL_587533\|20 | ···················· | ···················· | ···················· | ···················· | ···T················ | ···················· |
| M | 4161 | hCoV-19/England/QEUH-A1E1E4/2020\|EPI_ISL_587545\|20 | ···················· | ···················· | ···················· | ···················· | ·····T·············· | ···················· |
| M | 4162 | hCoV-19/England/QEUH-A218BC/2020\|EPI_ISL_587546\|20 | ···················· | ···················· | ···················· | ···················· | ···················· | ···················· |
| M | 4163 | hCoV-19/England/QEUH-A21FB7/2020\|EPI_ISL_587547\|20 | ···················· | ···················· | ···················· | ···················· | ···················· | ···················· |
| M | 4164 | hCoV-19/England/QEUH-A1E955/2020\|EPI_ISL_587548\|20 | ···················· | ···················· | ···················· | ···················· | ···················· | ···················· |
| M | 4165 | hCoV-19/England/QEUH-A1DFEB/2020\|EPI_ISL_587549\|20 | ···················· | ···················· | ···················· | ···················· | ···················· | ···················· |
| M | 4166 | hCoV-19/England/QEUH-A1DEDF/2020\|EPI_ISL_587551\|20 | ···················· | ···················· | ···················· | ···················· | ···················· | ···················· |
| M | 4167 | hCoV-19/England/QEUH-A1F453/2020\|EPI_ISL_587552\|20 | ···················· | ·A·················· | ···················· | ···················· | ···················· | ···················· |
| M | 4168 | hCoV-19/England/QEUH-A1EBB9/2020\|EPI_ISL_587553\|20 | ···················· | ···················· | ···················· | ···················· | ···················· | ···················· |
| M | 4169 | hCoV-19/England/QEUH-A1E15D/2020\|EPI_ISL_587554\|20 | ···················· | ···················· | ···················· | ···················· | ···················· | ···················· |
| M | 4170 | hCoV-19/England/QEUH-A1DF09/2020\|EPI_ISL_587555\|20 | ···················· | ···················· | ···················· | ···················· | ···················· | ···················· |
| M | 4171 | hCoV-19/England/QEUH-A1DC8A/2020\|EPI_ISL_587556\|20 | ···················· | ···················· | ···················· | ···················· | ···················· | ···················· |
| M | 4172 | hCoV-19/England/QEUH-A1E25A/2020\|EPI_ISL_587557\|20 | ···················· | ···················· | ···················· | ···················· | ···················· | ···················· |
| M | 4173 | hCoV-19/England/QEUH-A21F5D/2020\|EPI_ISL_587558\|20 | ···················· | ···················· | ···················· | ···················· | ···················· | ···················· |
| M | 4174 | hCoV-19/England/QEUH-A1DBD8/2020\|EPI_ISL_587559\|20 | ···················· | ···················· | ···················· | ···················· | ···················· | ···················· |
| M | 4175 | hCoV-19/England/QEUH-A22323/2020\|EPI_ISL_587560\|20 | ···················· | ···················· | ···················· | ···················· | ···T················ | ···················· |
| M | 4176 | hCoV-19/England/QEUH-A1DFCD/2020\|EPI_ISL_587561\|20 | ···················· | ···················· | ···················· | ···················· | ···················· | ···················· |
| M | 4177 | hCoV-19/England/QEUH-A1DF36/2020\|EPI_ISL_587562\|20 | ···················· | ···················· | ···················· | ···················· | ···················· | ···················· |
| M | 4178 | hCoV-19/England/QEUH-A1EAAD/2020\|EPI_ISL_587563\|20 | ···················· | ···················· | ···················· | ···················· | ···················· | ···················· |
| M | 4179 | hCoV-19/England/QEUH-A1E4CD/2020\|EPI_ISL_587564\|20 | ···················· | ···················· | ···················· | ···················· | ···················· | ···················· |
| M | 4180 | hCoV-19/England/QEUH-A1DE66/2020\|EPI_ISL_587565\|20 | ···················· | ···················· | ···················· | ···················· | ···················· | ···················· |
| M | 4181 | hCoV-19/England/QEUH-A1DE1B/2020\|EPI_ISL_587566\|20 | ···················· | ···················· | ···················· | ···················· | ···················· | ···················· |
| M | 4182 | hCoV-19/England/QEUH-A1F4F9/2020\|EPI_ISL_587567\|20 | ···················· | ···················· | ···················· | ···················· | ···················· | ···················· |
| M | 4183 | hCoV-19/England/QEUH-A2156A/2020\|EPI_ISL_587568\|20 | ···················· | ···················· | ···················· | ···················· | ···················· | ···················· |
| M | 4184 | hCoV-19/England/QEUH-A21904/2020\|EPI_ISL_587569\|20 | ···················· | ···················· | ···················· | ···················· | ···················· | ···················· |
| M | 4185 | hCoV-19/England/QEUH-A21342/2020\|EPI_ISL_587570\|20 | ···················· | ···················· | ···················· | ···················· | ···················· | ···················· |
| M | 4186 | hCoV-19/England/QEUH-A1DE2A/2020\|EPI_ISL_587571\|20 | ···················· | ···················· | ···················· | ···················· | ···················· | ···················· |
| M | 4187 | hCoV-19/England/QEUH-A1DF81/2020\|EPI_ISL_587572\|20 | ···················· | ···················· | ···················· | ···················· | ···················· | ···················· |
| M | 4188 | hCoV-19/England/QEUH-A1E894/2020\|EPI_ISL_587573\|20 | ···················· | ···················· | ···················· | ···················· | ···················· | ···················· |
| M | 4189 | hCoV-19/England/QEUH-A21290/2020\|EPI_ISL_587574\|20 | ···················· | ···················· | ···················· | ···················· | ···················· | ···················· |
| M | 4190 | hCoV-19/England/QEUH-A2162B/2020\|EPI_ISL_587575\|20 | ···················· | ···················· | ···················· | ···················· | ···················· | ···················· |
| M | 4191 | hCoV-19/England/QEUH-A210A5/2020\|EPI_ISL_587576\|20 | ···················· | ···················· | ···················· | ···················· | ···T················ | ···················· |
| M | 4192 | hCoV-19/England/QEUH-A224D5/2020\|EPI_ISL_587577\|20 | ···················· | ···················· | ···················· | ···················· | ···T················ | ···················· |
| M | 4193 | hCoV-19/England/QEUH-A2102D/2020\|EPI_ISL_587578\|20 | ···················· | ···················· | ···················· | ···················· | ···················· | ···················· |
| M | 4194 | hCoV-19/England/QEUH-A212AF/2020\|EPI_ISL_587579\|20 | ···················· | ···················· | ···················· | ···················· | ···················· | ···················· |
| M | 4195 | hCoV-19/England/QEUH-A202CE/2020\|EPI_ISL_587580\|20 | ···················· | ···················· | ···················· | ···················· | ···················· | ···················· |
| M | 4196 | hCoV-19/England/QEUH-A21263/2020\|EPI_ISL_587581\|20 | ···················· | ···················· | ···················· | ···················· | ···T················ | ···················· |
| M | 4197 | hCoV-19/England/QEUH-A21DEA/2020\|EPI_ISL_587582\|20 | ···················· | ···················· | ···················· | ···················· | ···················· | ···················· |
| M | 4198 | hCoV-19/England/QEUH-A1FCB5/2020\|EPI_ISL_587583\|20 | ···················· | ···················· | ···················· | ···················· | ···················· | ···················· |
| M | 4199 | hCoV-19/England/QEUH-A20932/2020\|EPI_ISL_587584\|20 | ···················· | ···················· | ···················· | ···················· | ···················· | ···················· |
| M | 4200 | hCoV-19/England/QEUH-A215D3/2020\|EPI_ISL_587585\|20 | ···················· | ···················· | ···················· | ···················· | ···T················ | ···················· |
| M | 4201 | hCoV-19/England/QEUH-A20307/2020\|EPI_ISL_587586\|20 | ···················· | ···················· | ···················· | ···················· | ···················· | ···················· |
| M | 4202 | hCoV-19/England/QEUH-A2248A/2020\|EPI_ISL_587587\|20 | ···················· | ···················· | ···················· | ···················· | ···················· | ···················· |
| M | 4203 | hCoV-19/England/QEUH-A215F1/2020\|EPI_ISL_587588\|20 | ···················· | ···················· | ···················· | ···················· | ···················· | ···················· |
| M | 4204 | hCoV-19/England/QEUH-A209C9/2020\|EPI_ISL_587589\|20 | ···················· | ···················· | ···················· | ···················· | ···················· | ···················· |
| M | 4205 | hCoV-19/England/QEUH-A1FA15/2020\|EPI_ISL_587591\|20 | ···················· | ···················· | ···················· | ···················· | ···················· | ···················· |
| M | 4206 | hCoV-19/England/QEUH-A2046E/2020\|EPI_ISL_587592\|20 | ···················· | ···················· | ···················· | ···················· | ···················· | ···················· |
| M | 4207 | hCoV-19/England/QEUH-A21069/2020\|EPI_ISL_587595\|20 | ···················· | ···················· | ···················· | ···················· | ···················· | ···················· |
| M | 4208 | hCoV-19/England/QEUH-A21E23/2020\|EPI_ISL_587596\|20 | ···················· | ···················· | ···················· | ···················· | ···················· | ···················· |
| M | 4209 | hCoV-19/England/QEUH-A20905/2020\|EPI_ISL_587597\|20 | ···················· | ···················· | ···················· | ···················· | ···················· | ···················· |
| M | 4210 | hCoV-19/England/QEUH-A22235/2020\|EPI_ISL_587598\|20 | ···················· | ···················· | ···················· | ···················· | ···················· | ···················· |
| M | 4211 | hCoV-19/England/QEUH-A1FA06/2020\|EPI_ISL_587599\|20 | ···················· | ···················· | ···················· | ···················· | ···················· | ···················· |
| M | 4212 | hCoV-19/England/QEUH-9F920B/2020\|EPI_ISL_588823\|20 | ···················· | ···················· | ···················· | ···················· | ···················· | ···················· |
| M | 4213 | hCoV-19/England/QEUH-9F6031/2020\|EPI_ISL_588829\|20 | ···················· | ···················· | ···················· | ···················· | ···················· | ···················· |
| M | 4214 | hCoV-19/England/QEUH-9F82C0/2020\|EPI_ISL_588843\|20 | ···················· | ···················· | ···················· | ···················· | ···················· | ···················· |
| M | 4215 | hCoV-19/England/QEUH-9F8C4A/2020\|EPI_ISL_588846\|20 | ···················· | ···················· | ···················· | ···················· | ···················· | ···················· |
| M | 4216 | hCoV-19/England/QEUH-9F9265/2020\|EPI_ISL_588847\|20 | ···················· | ···················· | ···················· | ···················· | ···················· | ···················· |
| M | 4217 | hCoV-19/England/QEUH-9F575B/2020\|EPI_ISL_588848\|20 | ···················· | ···················· | ···················· | ···················· | ···················· | ···················· |
| M | 4218 | hCoV-19/England/QEUH-9F6408/2020\|EPI_ISL_588849\|20 | ···················· | ···················· | ···················· | ···················· | ···················· | ···················· |
| M | 4219 | hCoV-19/England/QEUH-9F5885/2020\|EPI_ISL_588850\|20 | ···················· | ···················· | ···················· | ···················· | ···················· | ···················· |
| M | 4220 | hCoV-19/England/QEUH-9F8512/2020\|EPI_ISL_588851\|20 | ···················· | ···················· | ···················· | ···················· | ···················· | ···················· |
| M | 4221 | hCoV-19/England/QEUH-9F63DE/2020\|EPI_ISL_588852\|20 | ···················· | ···················· | ···················· | ···················· | ···················· | ···················· |
| M | 4222 | hCoV-19/England/QEUH-9F7C69/2020\|EPI_ISL_588853\|20 | ···················· | ···················· | ···················· | ···················· | ···················· | ···················· |
| M | 4223 | hCoV-19/England/QEUH-9F9317/2020\|EPI_ISL_588854\|20 | ···················· | ···················· | ···················· | ···················· | ···················· | ···················· |
| M | 4224 | hCoV-19/England/QEUH-9F81C3/2020\|EPI_ISL_588855\|20 | ···················· | ···················· | ···················· | ···················· | ···················· | ···················· |
| M | 4225 | hCoV-19/England/QEUH-9F7CD2/2020\|EPI_ISL_588856\|20 | ···················· | ···················· | ···················· | ···················· | ·····T·············· | ···················· |
| M | 4226 | hCoV-19/England/QEUH-9F7D1B/2020\|EPI_ISL_588857\|20 | ···················· | ···················· | ···················· | ···················· | ···················· | ···················· |
| M | 4227 | hCoV-19/England/QEUH-9F5ECF/2020\|EPI_ISL_588858\|20 | ···················· | ···················· | ···················· | ···················· | ···················· | ···················· |
| M | 4228 | hCoV-19/England/QEUH-9F82EE/2020\|EPI_ISL_588859\|20 | ···················· | ···················· | ···················· | ···················· | ···················· | ···················· |
| M | 4229 | hCoV-19/England/QEUH-9F60B9/2020\|EPI_ISL_588860\|20 | ···················· | ···················· | ···················· | ···················· | ···················· | ···················· |
| M | 4230 | hCoV-19/England/QEUH-9F6040/2020\|EPI_ISL_588861\|20 | ···················· | ···················· | ···················· | ···················· | ···················· | ···················· |
| M | 4231 | hCoV-19/England/QEUH-9F902F/2020\|EPI_ISL_588862\|20 | ···················· | ···················· | ···················· | ···················· | ···················· | ···················· |
| M | 4232 | hCoV-19/England/QEUH-9F8943/2020\|EPI_ISL_588863\|20 | ···················· | ···················· | ···················· | ···················· | ···················· | ···················· |
| M | 4233 | hCoV-19/England/QEUH-9F8C1D/2020\|EPI_ISL_588864\|20 | ···················· | ···················· | ···················· | ···················· | ···················· | ···················· |
| M | 4234 | hCoV-19/England/QEUH-9F92FC/2020\|EPI_ISL_588876\|20 | ···················· | ···················· | ···················· | ···················· | ···················· | ···················· |
| M | 4235 | hCoV-19/England/QEUH-9F863D/2020\|EPI_ISL_588877\|20 | ···················· | ···················· | ···················· | ···················· | ···················· | ···················· |
| M | 4236 | hCoV-19/England/QEUH-9F9344/2020\|EPI_ISL_588879\|20 | ···················· | ·A·················· | ···················· | ···················· | ···················· | ···················· |
| M | 4237 | hCoV-19/England/QEUH-9F903E/2020\|EPI_ISL_588880\|20 | ···················· | ···················· | ···················· | ···················· | ···················· | ···················· |
| M | 4238 | hCoV-19/England/QEUH-9F8CE0/2020\|EPI_ISL_588881\|20 | ···················· | ···················· | ···················· | ···················· | ···················· | ···················· |

M 4239 hCoV-19/England/QEUH-9F6329/2020|EPI_ISL_588883|20 ···················· ···················· ···················· ···················· ···················· ····················
M 4240 hCoV-19/England/QEUH-9F7CF0/2020|EPI_ISL_588884|20 ···················· ···················· ···················· ···················· ······T············· ····················
M 4241 hCoV-19/England/QEUH-9F8925/2020|EPI_ISL_588885|20 ···················· ···················· ···················· ···················· ···················· ····················
M 4242 hCoV-19/England/QEUH-9F62D1/2020|EPI_ISL_588886|20 ···················· ···················· ···················· ···················· ···················· ····················
M 4243 hCoV-19/England/QEUH-9F86D3/2020|EPI_ISL_588887|20 ···················· ···················· ···················· ···················· ····T··············· ····················
M 4244 hCoV-19/England/QEUH-9F63A1/2020|EPI_ISL_588888|20 ···················· ···················· ···················· ···················· ···················· ····················
M 4245 hCoV-19/England/QEUH-9F60E6/2020|EPI_ISL_588889|20 ···················· ···················· ···················· ···················· ···················· ····················
M 4246 hCoV-19/England/QEUH-9F64EA/2020|EPI_ISL_588890|20 ···················· ···················· ···················· ···················· ···················· ····················
M 4247 hCoV-19/England/QEUH-9F6453/2020|EPI_ISL_588891|20 ···················· ···················· ···················· ···················· ···················· ····················
M 4248 hCoV-19/England/QEUH-9F7CA5/2020|EPI_ISL_588892|20 ···················· ···················· ···················· ···················· ······T············· ····················
M 4249 hCoV-19/England/QEUH-9F8C95/2020|EPI_ISL_588893|20 ···················· ···················· ···················· ···················· ···················· ····················
M 4250 hCoV-19/England/QEUH-9F921A/2020|EPI_ISL_588894|20 ···················· ···················· ···················· ···················· ···················· ····················
M 4251 hCoV-19/England/QEUH-9F8E08/2020|EPI_ISL_588895|20 ···················· ···················· ···················· ···················· ···················· ····················
M 4252 hCoV-19/England/QEUH-9F6884/2020|EPI_ISL_588896|20 ···················· ···················· ···················· ···················· ···················· ····················
M 4253 hCoV-19/England/QEUH-9F6392/2020|EPI_ISL_588897|20 ···················· ···················· ···················· ···················· ···················· ····················
M 4254 hCoV-19/England/QEUH-9F8D38/2020|EPI_ISL_588898|20 ···················· ···················· ···················· ···················· ···················· ····················
M 4255 hCoV-19/England/QEUH-9F9247/2020|EPI_ISL_588899|20 ···················· ···················· ···················· ···················· ···················· ····················
M 4256 hCoV-19/Australia/VIC12549/2020|EPI_ISL_592022|202 ···················· ···················· ···················· ···················· ···················· ····················
M 4257 hCoV-19/Australia/VIC12552/2020|EPI_ISL_592023|202 ···················· ···················· ···················· ···················· ···················· ····················
M 4258 hCoV-19/Australia/VIC12565/2020|EPI_ISL_592024|202 ···················· ···················· ···················· ···················· ···················· ····················
M 4259 hCoV-19/Australia/VIC12573/2020|EPI_ISL_592025|202 ···················· ···················· ···················· ···················· ···················· ····················
M 4260 hCoV-19/Australia/VIC12578/2020|EPI_ISL_592026|202 ···················· ···················· ···················· ···················· ···················· ····················
M 4261 hCoV-19/Australia/VIC12580/2020|EPI_ISL_592027|202 ···················· ···················· ···················· ···················· ···················· ····················
M 4262 hCoV-19/Australia/VIC12583/2020|EPI_ISL_592028|202 ···················· ···················· ···················· ···················· ···················· ····················
M 4263 hCoV-19/Australia/VIC12587/2020|EPI_ISL_592029|202 ···················· ···················· ···················· ···················· ···················· ····················
M 4264 hCoV-19/Australia/VIC12588/2020|EPI_ISL_592030|202 ···················· ···················· ···················· ···················· ···················· ····················
M 4265 hCoV-19/Australia/VIC12594/2020|EPI_ISL_592031|202 ···················· ···················· ···················· ···················· ···················· ····················
M 4266 hCoV-19/Australia/VIC12596/2020|EPI_ISL_592032|202 ···················· ···················· ···················· ···················· ···················· ····················
M 4267 hCoV-19/Australia/VIC12600/2020|EPI_ISL_592033|202 ···················· ···················· ···················· ···················· ···················· ····················
M 4269 hCoV-19/Australia/VIC12603/2020|EPI_ISL_592035|202 ···················· ···················· ···················· ···················· ···················· ····················
M 4270 hCoV-19/Australia/VIC12606/2020|EPI_ISL_592036|202 ···················· ···················· ···················· ···················· ···················· ····················
M 4271 hCoV-19/Australia/VIC12612/2020|EPI_ISL_592037|202 ···················· ···················· ···················· ···················· ···················· ····················
M 4272 hCoV-19/Australia/VIC12613/2020|EPI_ISL_592038|202 ···················· ···················· ···················· ···················· ···················· ····················
M 4273 hCoV-19/Australia/VIC12615/2020|EPI_ISL_592039|202 ···················· ···················· ···················· ···················· ···················· ····················
M 4274 hCoV-19/Australia/VIC12616/2020|EPI_ISL_592040|202 ···················· ···················· ···················· ···················· ···················· ····················
M 4275 hCoV-19/Australia/VIC12639/2020|EPI_ISL_592041|202 ···················· ···················· ···················· ···················· ···················· ····················
M 4276 hCoV-19/Australia/VIC12643/2020|EPI_ISL_592042|202 ···················· ···················· ···················· ···················· ···················· ····················
M 4277 hCoV-19/Australia/VIC12647/2020|EPI_ISL_592043|202 ···················· ···················· ···················· ···················· ···················· ····················
M 4279 hCoV-19/Australia/VIC12651/2020|EPI_ISL_592045|202 ···················· ···················· ···················· ···················· ···················· ····················
M 4280 hCoV-19/Australia/VIC12653/2020|EPI_ISL_592046|202 ···················· ···················· ···················· ···················· ···················· ····················
M 4281 hCoV-19/Australia/VIC12658/2020|EPI_ISL_592047|202 ···················· ···················· ···················· ···················· ···················· ····················
M 4282 hCoV-19/Australia/VIC12660/2020|EPI_ISL_592048|202 ···················· ···················· ···················· ···················· ···················· ····················
M 4283 hCoV-19/Australia/VIC12668/2020|EPI_ISL_592049|202 ···················· ···················· ···················· ···················· ···················· ····················
M 4284 hCoV-19/Australia/VIC12670/2020|EPI_ISL_592050|202 ···················· ···················· ···················· ···················· ···················· ····················
M 4286 hCoV-19/Australia/VIC12672/2020|EPI_ISL_592052|202 ···················· ···················· ···················· ···················· ···················· ····················
M 4287 hCoV-19/Australia/VIC12679/2020|EPI_ISL_592053|202 ···················· ···················· ···················· ···················· ···················· ····················
M 4288 hCoV-19/Australia/VIC12682/2020|EPI_ISL_592054|202 ···················· ···················· ···················· ···················· ···················· ····················
M 4289 hCoV-19/Australia/VIC12684/2020|EPI_ISL_592055|202 ···················· ···················· ···················· ···················· ···················· ····················
M 4290 hCoV-19/Australia/VIC12689/2020|EPI_ISL_592056|202 ···················· ···················· ···················· ···················· ···················· ····················
M 4291 hCoV-19/Australia/VIC12692/2020|EPI_ISL_592057|202 ···················· ···················· ···················· ···················· ···················· ····················
M 4292 hCoV-19/Australia/VIC12698/2020|EPI_ISL_592058|202 ···················· ···················· ···················· ···················· ···················· ····················
M 4293 hCoV-19/Australia/VIC12700/2020|EPI_ISL_592059|202 ···················· ···················· ···················· ···················· ···················· ····················
M 4294 hCoV-19/Australia/VIC12701/2020|EPI_ISL_592060|202 ···················· ···················· ···················· ···················· ···················· ····················
M 4295 hCoV-19/Australia/VIC12721/2020|EPI_ISL_592061|202 ···················· ······A············· ···················· ···················· ···················· ····················
M 4296 hCoV-19/Australia/VIC12722/2020|EPI_ISL_592062|202 ···················· ···················· ···················· ···················· ···················· ····················
M 4297 hCoV-19/Australia/VIC12723/2020|EPI_ISL_592063|202 ···················· ···················· ···················· ···················· ···················· ····················
M 4298 hCoV-19/Australia/VIC12724/2020|EPI_ISL_592064|202 ···················· ···················· ···················· ···················· ···················· ····················
M 4299 hCoV-19/Australia/VIC12729/2020|EPI_ISL_592065|202 ···················· ···················· ···················· ···················· ···················· ····················
M 4301 hCoV-19/Australia/VIC12868/2020|EPI_ISL_592096|202 ···················· ···················· ···················· ···················· ···················· ····················
M 4302 hCoV-19/Australia/VIC15538/2020|EPI_ISL_593353|202 ···················· ···················· ···················· ···················· ···················· ····················
M 4303 hCoV-19/Australia/VIC15544/2020|EPI_ISL_593354|202 ···················· ···················· ···················· ···················· ···················· ····················
M 4304 hCoV-19/Australia/VIC15545/2020|EPI_ISL_593355|202 ···················· ···················· ···················· ···················· ···················· ····················
M 4305 hCoV-19/Australia/VIC15549/2020|EPI_ISL_593356|202 ···················· ···················· ···················· ···················· ···················· ····················
M 4306 hCoV-19/Australia/VIC15550/2020|EPI_ISL_593357|202 ···················· ···················· ···················· ···················· ···················· ····················
M 4308 hCoV-19/Australia/VIC15555/2020|EPI_ISL_593359|202 ···················· ···················· ···················· ···················· ···················· ····················
M 4309 hCoV-19/Australia/VIC15562/2020|EPI_ISL_593360|202 ···················· ···················· ···················· ···················· ···················· ····················
M 4310 hCoV-19/Australia/VIC15567/2020|EPI_ISL_593361|202 ···················· ···················· ···················· ···················· ···················· ····················
M 4311 hCoV-19/Australia/VIC15568/2020|EPI_ISL_593362|202 ···················· ···················· ···················· ···················· ···················· ····················
M 4312 hCoV-19/Australia/VIC15569/2020|EPI_ISL_593363|202 ···················· ···················· ···················· ···················· ···················· ····················
M 4313 hCoV-19/Australia/VIC15573/2020|EPI_ISL_593364|202 ···················· ···················· ···················· ···················· ···················· ····················
M 4314 hCoV-19/Australia/VIC15574/2020|EPI_ISL_593365|202 ···················· ···················· ···················· ···················· ···················· ····················
M 4315 hCoV-19/Australia/VIC15575/2020|EPI_ISL_593366|202 ···················· ···················· ···················· ···················· ···················· ····················
M 4316 hCoV-19/Australia/VIC15577/2020|EPI_ISL_593367|202 ···················· ···················· ···················· ···················· ···················· ····················
M 4317 hCoV-19/Australia/VIC15579/2020|EPI_ISL_593368|202 ···················· ···················· ···················· ···················· ···················· ····················
M 4320 hCoV-19/Australia/VIC15603/2020|EPI_ISL_593371|202 ···················· ···················· ···················· ···················· ···················· ····················
M 4321 hCoV-19/Australia/VIC15607/2020|EPI_ISL_593372|202 ···················· ···················· ···················· ···················· ···················· ····················
M 4322 hCoV-19/Australia/VIC15609/2020|EPI_ISL_593373|202 ···················· ···················· ···················· ···················· ···················· ····················
M 4323 hCoV-19/Australia/VIC15611/2020|EPI_ISL_593374|202 ···················· ···················· ···················· ···················· ···················· ····················
M 4324 hCoV-19/Australia/VIC15612/2020|EPI_ISL_593375|202 ···················· ···················· ···················· ···················· ···················· ····················
M 4326 hCoV-19/Australia/VIC15615/2020|EPI_ISL_593377|202 ···················· ···················· ···················· ···················· ··NN················ ····················

| | | | | | | | | |
|---|---|---|---|---|---|---|---|---|
| M | 4327 | hCoV-19/Australia/VIC15617/2020\|EPI_ISL_593378\|202 | ···················· | ···················· | ···················· | ···················· | ···················· | ···················· |
| M | 4328 | hCoV-19/Australia/VIC15619/2020\|EPI_ISL_593379\|202 | ···················· | ···················· | ···················· | ···················· | ···················· | ···················· |
| M | 4332 | hCoV-19/Australia/VIC15624/2020\|EPI_ISL_593383\|202 | ···················· | ···················· | ···················· | ···················· | ···················· | ···················· |
| M | 4333 | hCoV-19/Australia/VIC15625/2020\|EPI_ISL_593384\|202 | ···················· | ···················· | ···················· | ···················· | ···················· | ···················· |
| M | 4334 | hCoV-19/Australia/VIC15626/2020\|EPI_ISL_593385\|202 | ···················· | ···················· | ···················· | ···················· | ···················· | ···················· |
| M | 4335 | hCoV-19/Australia/VIC15627/2020\|EPI_ISL_593386\|202 | ···················· | ···················· | ···················· | ···················· | ···················· | ···················· |
| M | 4336 | hCoV-19/Australia/VIC15631/2020\|EPI_ISL_593387\|202 | ···················· | ···················· | ···················· | ···················· | ···················· | ···················· |
| M | 4337 | hCoV-19/Australia/VIC15633/2020\|EPI_ISL_593388\|202 | ···················· | ···················· | ···················· | ···················· | ···················· | ···················· |
| M | 4340 | hCoV-19/Australia/VIC15636/2020\|EPI_ISL_593391\|202 | ···················· | ···················· | ···················· | ···················· | ···················· | ···················· |
| M | 4341 | hCoV-19/Australia/VIC15639/2020\|EPI_ISL_593392\|202 | ···················· | ···················· | ···················· | ···················· | ···················· | ···················· |
| M | 4343 | hCoV-19/Australia/VIC15643/2020\|EPI_ISL_593394\|202 | ···················· | ···················· | ···················· | ···················· | ···················· | ···················· |
| M | 4346 | hCoV-19/Scotland/CVR4918/2020\|EPI_ISL_594684\|2020- | ···················· | ···················· | ···················· | ···················· | ···················· | ···················· |
| M | 4347 | hCoV-19/Scotland/CVR4921/2020\|EPI_ISL_594687\|2020- | ···················· | ·A·················· | ···················· | ···················· | ···················· | ···················· |
| M | 4348 | hCoV-19/Scotland/CVR4923/2020\|EPI_ISL_594689\|2020- | ···················· | ···················· | ···················· | ···················· | ···················· | ···················· |
| M | 4349 | hCoV-19/Scotland/CVR4926/2020\|EPI_ISL_594692\|2020- | ···················· | ···················· | ···················· | ···················· | ···················· | ···N················ |
| M | 4350 | hCoV-19/Scotland/CVR4927/2020\|EPI_ISL_594693\|2020- | ···················· | ···················· | ···················· | ···················· | ···················· | ···················· |
| M | 4351 | hCoV-19/Scotland/CVR4928/2020\|EPI_ISL_594694\|2020- | ···················· | ···················· | ···················· | ···················· | ···················· | ···················· |
| M | 4352 | hCoV-19/Scotland/CVR4930/2020\|EPI_ISL_594696\|2020- | ···················· | ···················· | ···················· | ···················· | ···················· | ···················· |
| M | 4353 | hCoV-19/Austria/CeMM0986/2020\|EPI_ISL_583830\|2020- | ···················· | ···················· | ···················· | ···················· | ···················· | ···················· |
| M | 4354 | hCoV-19/Austria/CeMM0987/2020\|EPI_ISL_583831\|2020- | ···················· | ···················· | ···················· | ···················· | ···················· | ···················· |
| M | 4355 | hCoV-19/Austria/CeMM0988/2020\|EPI_ISL_583832\|2020- | ···················· | ···················· | ···················· | ···················· | ···················· | ···················· |
| M | 4356 | hCoV-19/Austria/CeMM0989/2020\|EPI_ISL_583833\|2020- | ···················· | ···················· | ···················· | ···················· | ···················· | ···················· |
| M | 4357 | hCoV-19/Austria/CeMM0990/2020\|EPI_ISL_583834\|2020- | ···················· | ···················· | ···················· | ···················· | ···················· | ···················· |
| M | 4358 | hCoV-19/Austria/CeMM0991/2020\|EPI_ISL_583835\|2020- | ···················· | ···················· | ···················· | ···················· | ···················· | ···················· |
| M | 4359 | hCoV-19/Austria/CeMM0992/2020\|EPI_ISL_583836\|2020- | ···················· | ···················· | ···················· | ···················· | ···················· | ···················· |
| M | 4360 | hCoV-19/Austria/CeMM0993/2020\|EPI_ISL_583837\|2020- | ···················· | ···················· | ···················· | ···················· | ···················· | ···················· |
| M | 4361 | hCoV-19/Austria/CeMM0994/2020\|EPI_ISL_583838\|2020- | ···················· | ···················· | ···················· | ···················· | ···················· | ···················· |
| M | 4362 | hCoV-19/Austria/CeMM0995/2020\|EPI_ISL_583839\|2020- | ···················· | ···················· | ···················· | ···················· | ···················· | ···················· |
| M | 4363 | hCoV-19/Austria/CeMM0996/2020\|EPI_ISL_583840\|2020- | ···················· | ···················· | ···················· | ···················· | ···················· | ···················· |
| M | 4364 | hCoV-19/Austria/CeMM0997/2020\|EPI_ISL_583841\|2020- | ···················· | ···················· | ···················· | ···················· | ···················· | ···················· |
| M | 4365 | hCoV-19/Austria/CeMM0998/2020\|EPI_ISL_583842\|2020- | ···················· | ···················· | ···················· | ···················· | ···················· | ···················· |
| M | 4366 | hCoV-19/Austria/CeMM0999/2020\|EPI_ISL_583843\|2020- | ···················· | ···················· | ···················· | ···················· | ···················· | ···················· |
| M | 4367 | hCoV-19/Austria/CeMM1001/2020\|EPI_ISL_583844\|2020- | ···················· | ···················· | ···················· | ···················· | ···················· | ···················· |
| M | 4368 | hCoV-19/Austria/CeMM1004/2020\|EPI_ISL_583845\|2020- | ···················· | ···················· | ···················· | ···················· | ···················· | ···················· |
| M | 4369 | hCoV-19/Austria/CeMM1007/2020\|EPI_ISL_583847\|2020- | ···················· | ···················· | ···················· | ···················· | ···················· | ···················· |
| M | 4370 | hCoV-19/Austria/CeMM1008/2020\|EPI_ISL_583848\|2020- | ···················· | ···················· | ···················· | ···················· | ···················· | ···················· |
| M | 4371 | hCoV-19/Austria/CeMM1010/2020\|EPI_ISL_583849\|2020- | ···················· | ···················· | ···················· | ···················· | ···················· | ···················· |
| M | 4372 | hCoV-19/Austria/CeMM1011/2020\|EPI_ISL_583850\|2020- | ···················· | ···················· | ···················· | ···················· | ···················· | ···················· |
| M | 4373 | hCoV-19/Austria/CeMM1012/2020\|EPI_ISL_583851\|2020- | ···················· | ···················· | ···················· | ···················· | ···················· | ···················· |
| M | 4374 | hCoV-19/Austria/CeMM1013/2020\|EPI_ISL_583852\|2020- | ···················· | ···················· | ···················· | ···················· | ···················· | ···················· |
| M | 4375 | hCoV-19/Austria/CeMM1027/2020\|EPI_ISL_583853\|2020- | ···················· | ···················· | ···················· | ···················· | ···················· | ···················· |
| M | 4376 | hCoV-19/Austria/CeMM1031/2020\|EPI_ISL_583855\|2020- | ···················· | ···················· | ···················· | ···················· | ···················· | ···················· |
| M | 4377 | hCoV-19/Austria/CeMM1034/2020\|EPI_ISL_583857\|2020- | ···················· | ···················· | ···················· | ···················· | ···················· | ···················· |
| M | 4378 | hCoV-19/Austria/CeMM1035/2020\|EPI_ISL_583858\|2020- | ···················· | ···················· | ···················· | ···················· | ···················· | ···················· |
| M | 4379 | hCoV-19/Austria/CeMM1036/2020\|EPI_ISL_583859\|2020- | ···················· | ···················· | ···················· | ···················· | ···················· | ···················· |
| M | 4380 | hCoV-19/Austria/CeMM1043/2020\|EPI_ISL_583860\|2020- | ···················· | ···················· | ···················· | ···················· | ···················· | ···················· |
| M | 4381 | hCoV-19/Austria/CeMM1045/2020\|EPI_ISL_583862\|2020- | ···················· | ···················· | ···················· | ···················· | ···················· | ···················· |
| M | 4382 | hCoV-19/Austria/CeMM1046/2020\|EPI_ISL_583863\|2020- | ···················· | ···················· | ···················· | ···················· | ···················· | ···················· |
| M | 4383 | hCoV-19/Austria/CeMM1047/2020\|EPI_ISL_583864\|2020- | ···················· | ···················· | ···················· | ···················· | ···················· | ···················· |
| M | 4384 | hCoV-19/Austria/CeMM1048/2020\|EPI_ISL_583865\|2020- | ···················· | ···················· | ···················· | ···················· | ···················· | ···················· |
| M | 4385 | hCoV-19/Austria/CeMM1049/2020\|EPI_ISL_583866\|2020- | ···················· | ···················· | ···················· | ···················· | ···················· | ···················· |
| M | 4386 | hCoV-19/Austria/CeMM1055/2020\|EPI_ISL_583868\|2020- | ···················· | ···················· | ···················· | ···················· | ···················· | ···················· |
| M | 4387 | hCoV-19/Austria/CeMM1056/2020\|EPI_ISL_583869\|2020- | ···················· | ···················· | ···················· | ···················· | ···················· | ···················· |
| M | 4388 | hCoV-19/Austria/CeMM1057/2020\|EPI_ISL_583870\|2020- | ···················· | ···················· | ···················· | ···················· | ···················· | ···················· |
| M | 4389 | hCoV-19/Austria/CeMM1058/2020\|EPI_ISL_583871\|2020- | ···················· | ···················· | ···················· | ···················· | ···················· | ···················· |
| M | 4390 | hCoV-19/Austria/CeMM1059/2020\|EPI_ISL_583872\|2020- | ···················· | ···················· | ···················· | ···················· | ···················· | ···················· |
| M | 4391 | hCoV-19/Austria/CeMM1060/2020\|EPI_ISL_583873\|2020- | ···················· | ···················· | ···················· | ···················· | ···················· | ···················· |
| M | 4392 | hCoV-19/Austria/CeMM1061/2020\|EPI_ISL_583874\|2020- | ···················· | ···················· | ···················· | ···················· | ···················· | ···················· |
| M | 4393 | hCoV-19/Austria/CeMM1062/2020\|EPI_ISL_583875\|2020- | ···················· | ···················· | ···················· | ···················· | ···················· | ···················· |
| M | 4394 | hCoV-19/Austria/CeMM1063/2020\|EPI_ISL_583876\|2020- | ···················· | ···················· | ···················· | ···················· | ···················· | ···················· |
| M | 4395 | hCoV-19/Austria/CeMM1064/2020\|EPI_ISL_583877\|2020- | ···················· | ···················· | ···················· | ···················· | ···················· | ···················· |
| M | 4396 | hCoV-19/Austria/CeMM1065/2020\|EPI_ISL_583878\|2020- | ···················· | ···················· | ···················· | ···················· | ···················· | ···················· |
| M | 4397 | hCoV-19/Austria/CeMM1066/2020\|EPI_ISL_583879\|2020- | ···················· | ···················· | ···················· | ···················· | ···················· | ···················· |
| M | 4398 | hCoV-19/Austria/CeMM1067/2020\|EPI_ISL_583880\|2020- | ···················· | ···················· | ···················· | ···················· | ···················· | ···················· |
| M | 4399 | hCoV-19/Austria/CeMM1068/2020\|EPI_ISL_583881\|2020- | ···················· | ···················· | ···················· | ···················· | ···················· | ···················· |
| M | 4400 | hCoV-19/Austria/CeMM1077/2020\|EPI_ISL_583883\|2020- | ···········T········ | ···················· | ···················· | ···················· | ···················· | ···················· |
| M | 4401 | hCoV-19/Austria/CeMM1078/2020\|EPI_ISL_583884\|2020- | ···········T········ | ···················· | ···················· | ···················· | ···················· | ···················· |
| M | 4402 | hCoV-19/Austria/CeMM1083/2020\|EPI_ISL_583885\|2020- | ···················· | ·A·················· | ···················· | ···················· | ···················· | ···A················ |
| M | 4403 | hCoV-19/Austria/CeMM1084/2020\|EPI_ISL_583886\|2020- | ···················· | ···················· | ···················· | ···················· | ···················· | ···················· |
| M | 4404 | hCoV-19/Austria/CeMM1091/2020\|EPI_ISL_583887\|2020- | ···················· | ···················· | ···················· | ···················· | ···················· | ···················· |
| M | 4405 | hCoV-19/Austria/CeMM1092/2020\|EPI_ISL_583888\|2020- | ···················· | ···················· | ···················· | ···················· | ···················· | ···················· |
| M | 4406 | hCoV-19/Austria/CeMM1093/2020\|EPI_ISL_583889\|2020- | ···················· | ···················· | ···················· | ···················· | ···················· | ···················· |
| M | 4407 | hCoV-19/Austria/CeMM1094/2020\|EPI_ISL_583890\|2020- | ···················· | ···················· | ···················· | ···················· | ···················· | ···················· |
| M | 4408 | hCoV-19/Austria/CeMM1095/2020\|EPI_ISL_583891\|2020- | ···················· | ···················· | ···················· | ···················· | ···················· | ···················· |
| M | 4409 | hCoV-19/Singapore/1058/2020\|EPI_ISL_583893\|2020-09 | ···················· | ···················· | ···················· | ···················· | ···················· | ···················· |
| M | 4410 | hCoV-19/Singapore/1059/2020\|EPI_ISL_583894\|2020-08 | ···················· | ···················· | ···················· | ···················· | ···················· | ···················· |
| M | 4411 | hCoV-19/Singapore/1060/2020\|EPI_ISL_583895\|2020-08 | ···················· | ···················· | ···················· | ···················· | ···················· | ···················· |
| M | 4412 | hCoV-19/Singapore/1061/2020\|EPI_ISL_583896\|2020-08 | ···················· | ···················· | ···················· | ···················· | ···················· | ···················· |
| M | 4413 | hCoV-19/Singapore/1062/2020\|EPI_ISL_583897\|2020-08 | ···················· | ···················· | ···················· | ···················· | ···················· | ···················· |
| M | 4414 | hCoV-19/Singapore/1063/2020\|EPI_ISL_583898\|2020-08 | ···················· | ···················· | ···················· | ···················· | ···················· | ···················· |

| | | | | | | | | |
|---|---|---|---|---|---|---|---|---|
| M | 4415 | hCoV-19/Singapore/1064/2020\|EPI_ISL_583899\|2020-08 | ···················· | ···················· | ····················· | ···················· | ···················· | ···················· |
| M | 4416 | hCoV-19/England/QEUH-A22332/2020\|EPI_ISL_587500\|20 | ···················· | ···················· | ····················· | ···················· | ···················· | ···················· |
| M | 4417 | hCoV-19/England/QEUH-A1DF90/2020\|EPI_ISL_587501\|20 | ···················· | ···················· | ····················· | ···················· | ···················· | ···················· |
| M | 4418 | hCoV-19/England/QEUH-A1DA35/2020\|EPI_ISL_587502\|20 | ···················· | ···················· | ····················· | ···················· | ···················· | ···················· |
| M | 4419 | hCoV-19/Scotland/QEUH-A1F435/2020\|EPI_ISL_587503\|2 | ···················· | ···················· | ····················· | ···················· | ···················· | ···················· |
| M | 4420 | hCoV-19/England/QEUH-A1E9BF/2020\|EPI_ISL_587504\|20 | ···················· | ···················· | ····················· | ···················· | ···················· | ···················· |
| M | 4421 | hCoV-19/England/QEUH-A218F8/2020\|EPI_ISL_587505\|20 | ···················· | ···················· | ····················· | ···················· | ···················· | ···················· |
| M | 4422 | hCoV-19/England/QEUH-A1EB6E/2020\|EPI_ISL_587506\|20 | ···················· | ···················· | ····················· | ···················· | ···················· | ···················· |
| M | 4423 | hCoV-19/England/QEUH-A1E76A/2020\|EPI_ISL_587508\|20 | ···················· | ···················· | ····················· | ···················· | ···················· | ···················· |
| M | 4424 | hCoV-19/England/QEUH-A1DFAF/2020\|EPI_ISL_587509\|20 | ···················· | ···················· | ····················· | ···················· | ···················· | ···················· |
| M | 4425 | hCoV-19/England/QEUH-A1E296/2020\|EPI_ISL_587510\|20 | ···················· | ·A·················· | ····················· | ···················· | ···················· | ···A················ |
| M | 4426 | hCoV-19/England/QEUH-A1E014/2020\|EPI_ISL_587511\|20 | ···················· | ···················· | ····················· | ···················· | ···················· | ···················· |
| M | 4427 | hCoV-19/England/QEUH-9F83DC/2020\|EPI_ISL_588800\|20 | ···················· | ···················· | ····················· | ···················· | ···················· | ···················· |
| M | 4428 | hCoV-19/England/QEUH-9F89AD/2020\|EPI_ISL_588801\|20 | ···················· | ···················· | ····················· | ···················· | ···················· | ···················· |
| M | 4429 | hCoV-19/England/QEUH-9F9380/2020\|EPI_ISL_588802\|20 | ···················· | ···················· | ····················· | ···················· | ···················· | ···················· |
| M | 4430 | hCoV-19/England/QEUH-9F6383/2020\|EPI_ISL_588803\|20 | ···················· | ···················· | ····················· | ···················· | ···················· | ···················· |
| M | 4431 | hCoV-19/England/QEUH-9F92CF/2020\|EPI_ISL_588804\|20 | ···················· | ···················· | ····················· | ···················· | ····T··············· | ···················· |
| M | 4432 | hCoV-19/England/QEUH-9F7FD9/2020\|EPI_ISL_588805\|20 | ···················· | ···················· | ····················· | ···················· | ···················· | ···················· |
| M | 4433 | hCoV-19/England/QEUH-9F62A4/2020\|EPI_ISL_588806\|20 | ···················· | ···················· | ····················· | ···················· | ···················· | ···················· |
| M | 4434 | hCoV-19/England/QEUH-9F803F/2020\|EPI_ISL_588807\|20 | ···················· | ···················· | ····················· | ···················· | ···················· | ···················· |
| M | 4435 | hCoV-19/England/QEUH-9F92DE/2020\|EPI_ISL_588808\|20 | ···················· | ···················· | ····················· | ···················· | ···················· | ···················· |
| M | 4436 | hCoV-19/England/QEUH-9F63FC/2020\|EPI_ISL_588809\|20 | ···················· | ···················· | ····················· | ···················· | ···················· | ···················· |
| M | 4437 | hCoV-19/England/QEUH-9F8275/2020\|EPI_ISL_588810\|20 | ···················· | ···················· | ····················· | ···················· | ···················· | ···················· |
| M | 4438 | hCoV-19/England/QEUH-9F8B98/2020\|EPI_ISL_588812\|20 | ···················· | ···················· | ····················· | ···················· | ······T············· | ···················· |
| M | 4439 | hCoV-19/England/QEUH-9F7FF7/2020\|EPI_ISL_588813\|20 | ···················· | ···················· | ····················· | ···················· | ···················· | ···················· |
| M | 4440 | hCoV-19/England/QEUH-9F9326/2020\|EPI_ISL_588814\|20 | ···················· | ···················· | ····················· | ···················· | ···················· | ···················· |
| M | 4441 | hCoV-19/England/QEUH-9F7CB4/2020\|EPI_ISL_588815\|20 | ···················· | ···················· | ····················· | ···················· | ····T··············· | ···················· |
| M | 4442 | hCoV-19/England/QEUH-9F873A/2020\|EPI_ISL_588816\|20 | ···················· | ···················· | ····················· | ···················· | ···················· | ···················· |
| M | 4443 | hCoV-19/England/QEUH-9F8BF2/2020\|EPI_ISL_588817\|20 | ···················· | ···················· | ····················· | ···················· | ···················· | ···················· |
| M | 4444 | hCoV-19/England/QEUH-9F8E62/2020\|EPI_ISL_588818\|20 | ···················· | ···················· | ····················· | ···················· | ···················· | ···················· |
| M | 4445 | hCoV-19/England/QEUH-9F7FE8/2020\|EPI_ISL_588819\|20 | ···················· | ···················· | ····················· | ···················· | ···················· | ···················· |
| M | 4446 | hCoV-19/England/QEUH-9F7F60/2020\|EPI_ISL_588820\|20 | ···················· | ···················· | ····················· | ···················· | ···················· | ···················· |
| M | 4447 | hCoV-19/England/QEUH-9F56D6/2020\|EPI_ISL_588822\|20 | ···················· | ···················· | ····················· | ···················· | ···················· | ···················· |
| M | 4448 | hCoV-19/England/QEUH-9F8BD4/2020\|EPI_ISL_588825\|20 | ···················· | ···················· | ····················· | ···················· | ···················· | ···················· |
| M | 4449 | hCoV-19/England/QEUH-9F608C/2020\|EPI_ISL_588826\|20 | ···················· | ···················· | ····················· | ···················· | ···················· | ···················· |
| M | 4450 | hCoV-19/England/QEUH-9F63B0/2020\|EPI_ISL_588827\|20 | ···················· | ···················· | ····················· | ···················· | ···················· | ···················· |
| M | 4451 | hCoV-19/England/QEUH-9F8855/2020\|EPI_ISL_588828\|20 | ···················· | ···················· | ····················· | ···················· | ···················· | ···················· |
| M | 4452 | hCoV-19/England/QEUH-9F86C4/2020\|EPI_ISL_588830\|20 | ···················· | ···················· | ····················· | ···················· | ···················· | ···················· |
| M | 4453 | hCoV-19/England/QEUH-9F89CB/2020\|EPI_ISL_588831\|20 | ···················· | ·A·················· | ····················· | ···················· | ···················· | ···A················ |
| M | 4454 | hCoV-19/England/QEUH-9F85D6/2020\|EPI_ISL_588832\|20 | ···················· | ···················· | ····················· | ···················· | ···················· | ···················· |
| M | 4455 | hCoV-19/England/QEUH-9F8257/2020\|EPI_ISL_588833\|20 | ···················· | ···················· | ····················· | ···················· | ···················· | ···················· |
| M | 4456 | hCoV-19/England/QEUH-9F9238/2020\|EPI_ISL_588834\|20 | ···················· | ···················· | ····················· | ···················· | ···················· | ···················· |
| M | 4457 | hCoV-19/England/QEUH-9F82A2/2020\|EPI_ISL_588835\|20 | ···················· | ···················· | ····················· | ···················· | ···················· | ···················· |
| M | 4458 | hCoV-19/England/QEUH-9F6268/2020\|EPI_ISL_588836\|20 | ···················· | ···················· | ····················· | ···················· | ···················· | ···················· |
| M | 4459 | hCoV-19/England/QEUH-9F8B89/2020\|EPI_ISL_588837\|20 | ···················· | ···················· | ····················· | ···················· | ···················· | ···················· |
| M | 4460 | hCoV-19/England/QEUH-9F8776/2020\|EPI_ISL_588838\|20 | ···················· | ···················· | ····················· | ···················· | ···················· | ···················· |
| M | 4461 | hCoV-19/England/QEUH-9F8BE3/2020\|EPI_ISL_588839\|20 | ···················· | ···················· | ····················· | ···················· | ······T············· | ···················· |
| M | 4462 | hCoV-19/England/QEUH-9F85E5/2020\|EPI_ISL_588840\|20 | ···················· | ···················· | ····················· | ···················· | ···················· | ···················· |
| M | 4463 | hCoV-19/England/QEUH-9F8794/2020\|EPI_ISL_588841\|20 | ···················· | ···················· | ····················· | ···················· | ···················· | ···················· |
| M | 4464 | hCoV-19/England/QEUH-9F57E2/2020\|EPI_ISL_588842\|20 | ···················· | ···················· | ····················· | ···················· | ···················· | ···················· |
| M | 4465 | hCoV-19/Australia/VIC12730/2020\|EPI_ISL_592066\|202 | ···················· | ···················· | ····················· | ···················· | ···················· | ···················· |
| M | 4467 | hCoV-19/Australia/VIC12736/2020\|EPI_ISL_592068\|202 | ···················· | ···················· | ····················· | ···················· | ···················· | ···················· |
| M | 4469 | hCoV-19/Australia/VIC12740/2020\|EPI_ISL_592070\|202 | ···················· | ···················· | ····················· | ···················· | ···················· | ···················· |
| M | 4470 | hCoV-19/Australia/VIC12757/2020\|EPI_ISL_592071\|202 | ···················· | ···················· | ····················· | ···················· | ···················· | ···················· |
| M | 4472 | hCoV-19/Australia/VIC12761/2020\|EPI_ISL_592073\|202 | ···················· | ···················· | ····················· | ···················· | ···················· | ···················· |
| M | 4473 | hCoV-19/Australia/VIC12762/2020\|EPI_ISL_592074\|202 | ···················· | ···················· | ····················· | ···················· | ···················· | ···················· |
| M | 4474 | hCoV-19/Australia/VIC12764/2020\|EPI_ISL_592075\|202 | ···················· | ···················· | ····················· | ···················· | ···················· | ···················· |
| M | 4475 | hCoV-19/Australia/VIC12765/2020\|EPI_ISL_592076\|202 | ···················· | ···················· | ····················· | ···················· | ···················· | ···················· |
| M | 4476 | hCoV-19/Australia/VIC12778/2020\|EPI_ISL_592077\|202 | ···················· | ···················· | ····················· | ···················· | ···················· | ···················· |
| M | 4478 | hCoV-19/Australia/VIC12787/2020\|EPI_ISL_592079\|202 | ···················· | ···················· | ····················· | ···················· | ···················· | ···R················ |
| M | 4479 | hCoV-19/Australia/VIC12790/2020\|EPI_ISL_592080\|202 | ···················· | ···················· | ····················· | ···················· | ···················· | ···················· |
| M | 4481 | hCoV-19/Australia/VIC12801/2020\|EPI_ISL_592082\|202 | ···················· | ···················· | ····················· | ···················· | ···················· | ···················· |
| M | 4482 | hCoV-19/Australia/VIC12802/2020\|EPI_ISL_592083\|202 | ···················· | ···················· | ····················· | ···················· | ···················· | ···················· |
| M | 4483 | hCoV-19/Australia/VIC12810/2020\|EPI_ISL_592084\|202 | ···················· | ···················· | ····················· | ···················· | ···················· | ···················· |
| M | 4484 | hCoV-19/Australia/VIC12825/2020\|EPI_ISL_592085\|202 | ···················· | ···················· | ····················· | ···················· | ···················· | ···················· |
| M | 4485 | hCoV-19/Australia/VIC12826/2020\|EPI_ISL_592086\|202 | ···················· | ···················· | ····················· | ···················· | ···················· | ···················· |
| M | 4486 | hCoV-19/Australia/VIC12831/2020\|EPI_ISL_592087\|202 | ···················· | ···················· | ····················· | ···················· | ···················· | ···················· |
| M | 4487 | hCoV-19/Australia/VIC12832/2020\|EPI_ISL_592088\|202 | ···················· | ······R············· | ····················· | ···················· | ···················· | ···················· |
| M | 4488 | hCoV-19/Australia/VIC12837/2020\|EPI_ISL_592089\|202 | ···················· | ···················· | ····················· | ···················· | ···················· | ···················· |
| M | 4489 | hCoV-19/Australia/VIC12858/2020\|EPI_ISL_592091\|202 | ···················· | ···················· | ····················· | ···················· | ···················· | ···················· |
| M | 4490 | hCoV-19/Australia/VIC12863/2020\|EPI_ISL_592092\|202 | ···················· | ···················· | ····················· | ···················· | ···················· | ···················· |
| M | 4491 | hCoV-19/Australia/VIC12864/2020\|EPI_ISL_592093\|202 | ···················· | ···················· | ····················· | ···················· | ···················· | ···················· |
| M | 4492 | hCoV-19/Australia/VIC12866/2020\|EPI_ISL_592094\|202 | ···················· | ···················· | ····················· | ···················· | ···················· | ···················· |
| M | 4493 | hCoV-19/Australia/VIC12867/2020\|EPI_ISL_592095\|202 | ···················· | ···················· | ····················· | ···················· | ···················· | ···················· |
| M | 4494 | hCoV-19/Australia/VIC12869/2020\|EPI_ISL_592097\|202 | ···················· | ···················· | ····················· | ···················· | ···················· | ···················· |
| M | 4495 | hCoV-19/Australia/VIC12871/2020\|EPI_ISL_592098\|202 | ···················· | ···················· | ····················· | ···················· | ···················· | ···················· |
| M | 4497 | hCoV-19/Australia/VIC15223/2020\|EPI_ISL_593201\|202 | ···················· | ···················· | ····················· | ···················· | ···················· | ···················· |
| M | 4498 | hCoV-19/Australia/VIC15225/2020\|EPI_ISL_593202\|202 | ···················· | ···················· | ····················· | ···················· | ···················· | ···················· |
| M | 4499 | hCoV-19/Australia/VIC15226/2020\|EPI_ISL_593203\|202 | ···················· | ···················· | ····················· | ···················· | ···················· | ···················· |
| M | 4500 | hCoV-19/Australia/VIC15227/2020\|EPI_ISL_593204\|202 | ···················· | ···················· | ····················· | ···················· | ···················· | ···················· |

| | | | | | | | | |
|---|---|---|---|---|---|---|---|---|
| M | 4501 | hCoV-19/Australia/VIC15229/2020\|EPI_ISL_593205\|202 | ···················· | ···················· | ···················· | ···················· | ···················· | ···················· |
| M | 4502 | hCoV-19/Australia/VIC15230/2020\|EPI_ISL_593206\|202 | ···················· | ···················· | ···················· | ···················· | ···················· | ···················· |
| M | 4503 | hCoV-19/Australia/VIC15235/2020\|EPI_ISL_593210\|202 | ···················· | ···················· | ···················· | ···················· | ···················· | ···················· |
| M | 4504 | hCoV-19/Australia/VIC15237/2020\|EPI_ISL_593211\|202 | ···················· | ···················· | ···················· | ···················· | ···················· | ···················· |
| M | 4505 | hCoV-19/Australia/VIC15239/2020\|EPI_ISL_593212\|202 | ···················· | ···················· | ···················· | ···················· | ···················· | ···················· |
| M | 4507 | hCoV-19/Australia/VIC15241/2020\|EPI_ISL_593214\|202 | ···················· | ···················· | ···················· | ···················· | ···················· | ···················· |
| M | 4508 | hCoV-19/Australia/VIC15242/2020\|EPI_ISL_593215\|202 | ···················· | ···················· | ···················· | ···················· | ···················· | ···················· |
| M | 4510 | hCoV-19/Australia/VIC15245/2020\|EPI_ISL_593217\|202 | ···················· | ···················· | ···················· | ···················· | ···················· | ···················· |
| M | 4511 | hCoV-19/Australia/VIC15249/2020\|EPI_ISL_593218\|202 | ···················· | ···················· | ···················· | ···················· | ···················· | ···················· |
| M | 4512 | hCoV-19/Australia/VIC15250/2020\|EPI_ISL_593219\|202 | ···················· | ···················· | ···················· | ···················· | ···················· | ···················· |
| M | 4514 | hCoV-19/Australia/VIC15256/2020\|EPI_ISL_593221\|202 | ···················· | ···················· | ···················· | ···················· | ···················· | ···················· |
| M | 4515 | hCoV-19/Australia/VIC15257/2020\|EPI_ISL_593222\|202 | ···················· | ···················· | ···················· | ···················· | ···················· | ·········R·········· |
| M | 4516 | hCoV-19/Australia/VIC15261/2020\|EPI_ISL_593223\|202 | ···················· | ···················· | ···················· | ···················· | ···················· | ···················· |
| M | 4517 | hCoV-19/Australia/VIC15265/2020\|EPI_ISL_593224\|202 | ···················· | ···················· | ···················· | ···················· | ···················· | ···················· |
| M | 4518 | hCoV-19/Australia/VIC15268/2020\|EPI_ISL_593225\|202 | ···················· | ···················· | ···················· | ···················· | ···················· | ···················· |
| M | 4519 | hCoV-19/Australia/VIC15269/2020\|EPI_ISL_593226\|202 | ···················· | ···················· | ···················· | ···················· | ···················· | ···················· |
| M | 4520 | hCoV-19/Australia/VIC15270/2020\|EPI_ISL_593227\|202 | ···················· | ···················· | ···················· | ···················· | ···················· | ···················· |
| M | 4521 | hCoV-19/Australia/VIC15271/2020\|EPI_ISL_593228\|202 | ···················· | ···················· | ···················· | ···················· | ···················· | ···················· |
| M | 4522 | hCoV-19/Australia/VIC15272/2020\|EPI_ISL_593229\|202 | ···················· | ···················· | ···················· | ···················· | ···················· | ···················· |
| M | 4523 | hCoV-19/Australia/VIC15276/2020\|EPI_ISL_593230\|202 | ···················· | ···················· | ···················· | ···················· | ···················· | ···················· |
| M | 4524 | hCoV-19/Australia/VIC15277/2020\|EPI_ISL_593231\|202 | ···················· | ···················· | ···················· | ···················· | ···················· | ···················· |
| M | 4525 | hCoV-19/Australia/VIC15654/2020\|EPI_ISL_593397\|202 | ···················· | ···················· | ···················· | ···················· | ···················· | ···················· |
| M | 4529 | hCoV-19/England/BIRM-64671/2020\|EPI_ISL_594502\|202 | ···················· | ·A·················· | ···················· | ···················· | ···················· | ··A················· |
| M | 4530 | hCoV-19/England/BIRM-646DB/2020\|EPI_ISL_594506\|202 | ···················· | ·A·················· | ···················· | ···················· | ···················· | ··A················· |
| M | 4531 | hCoV-19/England/BIRM-64723/2020\|EPI_ISL_594510\|202 | ···················· | ···················· | ···················· | ···················· | ···················· | ···················· |
| M | 4532 | hCoV-19/England/BIRM-64732/2020\|EPI_ISL_594511\|202 | ···················· | ·A·················· | ···················· | ···················· | ···················· | ··A················· |
| M | 4533 | hCoV-19/England/BIRM-6477E/2020\|EPI_ISL_594512\|202 | ···················· | ···················· | ···················· | ···················· | ···················· | ···················· |
| M | 4534 | hCoV-19/England/BIRM-6478D/2020\|EPI_ISL_594513\|202 | ···················· | ···················· | ···················· | ···················· | ···················· | ···················· |
| M | 4537 | hCoV-19/England/BIRM-64E5B/2020\|EPI_ISL_594523\|202 | ···················· | ·A·················· | ···················· | ···················· | ···················· | ··A················· |
| M | 4539 | hCoV-19/England/BIRM-64E79/2020\|EPI_ISL_594525\|202 | ···················· | ···················· | ···················· | ···················· | ···················· | ···················· |
| M | 4540 | hCoV-19/England/BIRM-64E97/2020\|EPI_ISL_594526\|202 | ···················· | ·A·················· | ···················· | ···················· | ···················· | ··A················· |
| M | 4541 | hCoV-19/England/BIRM-64EB5/2020\|EPI_ISL_594527\|202 | ···················· | ·A·················· | ···················· | ···················· | ···················· | ··A················· |
| M | 4542 | hCoV-19/England/BIRM-64EC4/2020\|EPI_ISL_594528\|202 | ···················· | ···················· | ···················· | ···················· | ···················· | ···················· |
| M | 4543 | hCoV-19/England/BIRM-64EE2/2020\|EPI_ISL_594529\|202 | ···················· | ···················· | ···················· | ···················· | ···················· | ···················· |
| M | 4544 | hCoV-19/England/BIRM-64F2B/2020\|EPI_ISL_594532\|202 | ···················· | ···················· | ···················· | ···················· | ···················· | ···················· |
| M | 4545 | hCoV-19/England/BIRM-64F3A/2020\|EPI_ISL_594533\|202 | ···················· | ···················· | ···················· | ···················· | ···················· | ···················· |
| M | 4549 | hCoV-19/England/BIRM-65027/2020\|EPI_ISL_594542\|202 | ···················· | ···················· | ···················· | ···················· | ···················· | ···················· |
| M | 4550 | hCoV-19/England/BIRM-6527C/2020\|EPI_ISL_594543\|202 | ···················· | ···················· | ···················· | ···················· | ···················· | ···················· |
| M | 4551 | hCoV-19/England/BIRM-6529A/2020\|EPI_ISL_594544\|202 | ···················· | ···················· | ···················· | ···················· | ···················· | ···················· |
| M | 4553 | hCoV-19/England/BIRM-6531F/2020\|EPI_ISL_594547\|202 | ···················· | ···················· | ···················· | ···················· | ···················· | ···················· |
| M | 4554 | hCoV-19/England/BIRM-6534C/2020\|EPI_ISL_594550\|202 | ···················· | ···················· | ···················· | ···················· | ···················· | ···················· |
| M | 4555 | hCoV-19/England/BIRM-65388/2020\|EPI_ISL_594551\|202 | ···················· | ·A·················· | ···················· | ···················· | ···················· | ··A················· |
| M | 4556 | hCoV-19/England/BIRM-653A6/2020\|EPI_ISL_594552\|202 | ···················· | ···················· | ···················· | ···················· | ···················· | ···················· |
| M | 4557 | hCoV-19/England/BIRM-653B5/2020\|EPI_ISL_594553\|202 | ···················· | ···················· | ···················· | ···················· | ···················· | ···················· |
| M | 4558 | hCoV-19/England/BIRM-653D3/2020\|EPI_ISL_594555\|202 | ···················· | ···················· | ···················· | ···················· | ···················· | ···················· |
| M | 4559 | hCoV-19/England/BIRM-653E2/2020\|EPI_ISL_594556\|202 | ···················· | ···················· | ···················· | ···················· | ···················· | ···················· |
| M | 4560 | hCoV-19/England/BIRM-653F1/2020\|EPI_ISL_594557\|202 | ···················· | ···················· | ···················· | ···················· | ···················· | ···················· |
| M | 4563 | hCoV-19/Scotland/QEUH-A013FD/2020\|EPI_ISL_595811\|2 | ···················· | ···················· | ···················· | ···················· | ···················· | ···················· |
| M | 4564 | hCoV-19/England/QEUH-A01445/2020\|EPI_ISL_595813\|20 | ···················· | ···················· | ···················· | ···················· | ···················· | ···················· |
| M | 4565 | hCoV-19/England/QEUH-A01454/2020\|EPI_ISL_595814\|20 | ···················· | ···················· | ···················· | ···················· | ···················· | ···················· |
| M | 4566 | hCoV-19/England/QEUH-A016E5/2020\|EPI_ISL_595816\|20 | ···················· | ···················· | ···················· | ···················· | ···················· | ···················· |
| M | 4567 | hCoV-19/England/QEUH-A016F4/2020\|EPI_ISL_595817\|20 | ···················· | ···················· | ···················· | ···················· | ···················· | ···················· |
| M | 4568 | hCoV-19/England/QEUH-A01ACB/2020\|EPI_ISL_595824\|20 | ···················· | ···················· | ···················· | ···················· | ···················· | ···················· |
| M | 4569 | hCoV-19/England/QEUH-A01ADA/2020\|EPI_ISL_595825\|20 | ···················· | ···················· | ···················· | ···················· | ···················· | ···················· |
| M | 4570 | hCoV-19/England/QEUH-A01B9B/2020\|EPI_ISL_595826\|20 | ···················· | ·A·················· | ···················· | ···················· | ···················· | ···················· |
| M | 4571 | hCoV-19/Scotland/QEUH-A01C01/2020\|EPI_ISL_595828\|2 | ···················· | ···················· | ···················· | ···················· | ···················· | ···················· |
| M | 4572 | hCoV-19/England/QEUH-A01C10/2020\|EPI_ISL_595829\|20 | ···················· | ···················· | ···················· | ···················· | ···················· | ···················· |
| M | 4573 | hCoV-19/England/QEUH-A01C4D/2020\|EPI_ISL_595831\|20 | ···················· | ···················· | ···················· | ···················· | ···················· | ···················· |
| M | 4574 | hCoV-19/England/QEUH-A01C6B/2020\|EPI_ISL_595832\|20 | ···················· | ···················· | ···················· | ···················· | ···················· | ···················· |
| M | 4575 | hCoV-19/England/QEUH-A01C7A/2020\|EPI_ISL_595833\|20 | ···················· | ···················· | ···················· | ···················· | ···················· | ···················· |
| M | 4576 | hCoV-19/Scotland/QEUH-A01C98/2020\|EPI_ISL_595834\|2 | ···················· | ···················· | ···················· | ···················· | ···················· | ···················· |
| M | 4577 | hCoV-19/England/QEUH-A01CA7/2020\|EPI_ISL_595835\|20 | ···················· | ···················· | ···················· | ···················· | ···················· | ···················· |
| M | 4578 | hCoV-19/Scotland/QEUH-A02936/2020\|EPI_ISL_595843\|2 | ···················· | ···················· | ···················· | ···················· | ···················· | ···················· |
| M | 4582 | hCoV-19/England/QEUH-A3100C/2020\|EPI_ISL_595852\|20 | ···················· | ···················· | ···················· | ···················· | ···················· | ···················· |
| M | 4583 | hCoV-19/England/QEUH-A3102A/2020\|EPI_ISL_595854\|20 | ···················· | ···················· | ···················· | ···················· | ···················· | ···················· |
| M | 4585 | hCoV-19/England/QEUH-A31048/2020\|EPI_ISL_595856\|20 | ···················· | ···················· | ···················· | ···················· | ···················· | ···················· |
| M | 4587 | hCoV-19/England/QEUH-A310B1/2020\|EPI_ISL_595862\|20 | ···················· | ···················· | ···················· | ···················· | ···················· | ···················· |
| M | 4588 | hCoV-19/England/QEUH-A310DF/2020\|EPI_ISL_595863\|20 | ···················· | ···················· | ···················· | ···················· | ·····T·············· | ···················· |
| M | 4589 | hCoV-19/England/QEUH-A310EE/2020\|EPI_ISL_595864\|20 | ···················· | ···················· | ···················· | ···················· | ···················· | ···················· |
| M | 4591 | hCoV-19/England/QEUH-A31127/2020\|EPI_ISL_595867\|20 | ···················· | ···················· | ···················· | ···················· | ···················· | ···················· |
| M | 4592 | hCoV-19/England/QEUH-A31172/2020\|EPI_ISL_595871\|20 | ···················· | ···················· | ···················· | ···················· | ···················· | ···················· |
| M | 4593 | hCoV-19/England/QEUH-A31242/2020\|EPI_ISL_595872\|20 | ···················· | ···················· | ···················· | ···················· | ···················· | ···················· |
| M | 4594 | hCoV-19/England/QEUH-A3127F/2020\|EPI_ISL_595875\|20 | ···················· | ···················· | ···················· | ···················· | ···················· | ···················· |
| M | 4596 | hCoV-19/England/QEUH-A3129D/2020\|EPI_ISL_595877\|20 | ···················· | ···················· | ···················· | ···················· | ···················· | ···················· |
| M | 4597 | hCoV-19/England/QEUH-A31312/2020\|EPI_ISL_595878\|20 | ···················· | ···················· | ···················· | ···················· | ···················· | ···················· |
| M | 4598 | hCoV-19/England/QEUH-A31321/2020\|EPI_ISL_595879\|20 | ···················· | ···················· | ···················· | ···················· | ···················· | ···················· |
| M | 4599 | hCoV-19/England/QEUH-A3134F/2020\|EPI_ISL_595880\|20 | ···················· | ···················· | ···················· | ···················· | ···················· | ···················· |
| M | 4600 | hCoV-19/England/QEUH-A3138B/2020\|EPI_ISL_595882\|20 | ···················· | ···················· | ···················· | ···················· | ···················· | ···················· |
| M | 4601 | hCoV-19/England/QEUH-A313B8/2020\|EPI_ISL_595884\|20 | ···················· | ···················· | ···················· | ···················· | ···················· | ···················· |
| M | 4602 | hCoV-19/England/QEUH-A3141F/2020\|EPI_ISL_595886\|20 | ···················· | ···················· | ···················· | ···················· | ···················· | ···················· |

| | | | | | | | | | |
|---|---|---|---|---|---|---|---|---|---|
| M | 4603 | hCoV-19/England/QEUH-A3144C/2020\|EPI_ISL_595888\|20 | ···················· | ···················· | ···················· | ···················· | ···················· | ···················· | |
| M | 4604 | hCoV-19/England/QEUH-A3145B/2020\|EPI_ISL_595889\|20 | ···················· | ···················· | ···················· | ···················· | ···················· | ···················· | |
| M | 4605 | hCoV-19/England/QEUH-A3146A/2020\|EPI_ISL_595890\|20 | ···················· | ···················· | ···················· | ···················· | ···················· | ···················· | |
| M | 4606 | hCoV-19/England/QEUH-A31479/2020\|EPI_ISL_595891\|20 | ···················· | ···················· | ···················· | ···················· | ···················· | ···················· | |
| M | 4607 | hCoV-19/England/QEUH-A31488/2020\|EPI_ISL_595892\|20 | ···················· | ···················· | ···················· | ···················· | ···················· | ···················· | |
| M | 4608 | hCoV-19/England/QEUH-A31497/2020\|EPI_ISL_595893\|20 | ···················· | ···················· | ···················· | ···················· | ···················· | ···················· | |
| M | 4609 | hCoV-19/Austria/CeMM0948/2020\|EPI_ISL_583800\|2020- | ···················· | ···················· | ···················· | ···················· | ···················· | ···················· | |
| M | 4610 | hCoV-19/Austria/CeMM0949/2020\|EPI_ISL_583801\|2020- | ···················· | ···················· | ···················· | ···················· | ···················· | ···················· | |
| M | 4611 | hCoV-19/Austria/CeMM0950/2020\|EPI_ISL_583802\|2020- | ···················· | ···················· | ···················· | ···················· | ···················· | ···················· | |
| M | 4612 | hCoV-19/Austria/CeMM0953/2020\|EPI_ISL_583803\|2020- | ···················· | ···················· | ···················· | ···················· | ···················· | ···················· | |
| M | 4613 | hCoV-19/Austria/CeMM0954/2020\|EPI_ISL_583804\|2020- | ···················· | ···················· | ···················· | ···················· | ···················· | ···················· | |
| M | 4614 | hCoV-19/Austria/CeMM0956/2020\|EPI_ISL_583805\|2020- | ···················· | ···················· | ···················· | ···················· | ···················· | ···················· | |
| M | 4615 | hCoV-19/Austria/CeMM0957/2020\|EPI_ISL_583806\|2020- | ···················· | ···················· | ···················· | ···················· | ···················· | ···················· | |
| M | 4616 | hCoV-19/Austria/CeMM0958/2020\|EPI_ISL_583807\|2020- | ···················· | ···················· | ···················· | ···················· | ···················· | ···················· | |
| M | 4617 | hCoV-19/Austria/CeMM0959/2020\|EPI_ISL_583808\|2020- | ···················· | ···················· | ···················· | ···················· | ···················· | ···················· | |
| M | 4618 | hCoV-19/Austria/CeMM0960/2020\|EPI_ISL_583809\|2020- | ···················· | ···················· | ···················· | ···················· | ···················· | ···················· | |
| M | 4619 | hCoV-19/Austria/CeMM0961/2020\|EPI_ISL_583810\|2020- | ···················· | ···················· | ···················· | ···················· | ···················· | ···················· | |
| M | 4620 | hCoV-19/Austria/CeMM0962/2020\|EPI_ISL_583811\|2020- | ···················· | ···················· | ···················· | ···················· | ···················· | ···················· | |
| M | 4621 | hCoV-19/Austria/CeMM0963/2020\|EPI_ISL_583812\|2020- | ···················· | ···················· | ···················· | ···················· | ···················· | ···················· | |
| M | 4622 | hCoV-19/Austria/CeMM0964/2020\|EPI_ISL_583813\|2020- | ···················· | ···················· | ···················· | ···················· | ···················· | ···················· | |
| M | 4623 | hCoV-19/Austria/CeMM0965/2020\|EPI_ISL_583814\|2020- | ···················· | ···················· | ···················· | ···················· | ···················· | ···················· | |
| M | 4624 | hCoV-19/Austria/CeMM0966/2020\|EPI_ISL_583815\|2020- | ···················· | ···················· | ···················· | ···················· | ···················· | ···················· | |
| M | 4625 | hCoV-19/Austria/CeMM0970/2020\|EPI_ISL_583816\|2020- | ···················· | ···················· | ···················· | ···················· | ···················· | ···················· | |
| M | 4626 | hCoV-19/Austria/CeMM0971/2020\|EPI_ISL_583817\|2020- | ···················· | ···················· | ···················· | ···················· | ···················· | ···················· | |
| M | 4627 | hCoV-19/Austria/CeMM0972/2020\|EPI_ISL_583818\|2020- | ···················· | ···················· | ···················· | ···················· | ···················· | ···················· | |
| M | 4628 | hCoV-19/Austria/CeMM0973/2020\|EPI_ISL_583819\|2020- | ···················· | ···················· | ···················· | ···················· | ···················· | ···················· | |
| M | 4629 | hCoV-19/Austria/CeMM0975/2020\|EPI_ISL_583820\|2020- | ···················· | ···················· | ···················· | ···················· | ···················· | ···················· | |
| M | 4630 | hCoV-19/Austria/CeMM0976/2020\|EPI_ISL_583821\|2020- | ···················· | ···················· | ···················· | ···················· | ···················· | ···················· | |
| M | 4631 | hCoV-19/Austria/CeMM0977/2020\|EPI_ISL_583822\|2020- | ···················· | ···················· | ···················· | ···················· | ···················· | ···················· | |
| M | 4632 | hCoV-19/Austria/CeMM0978/2020\|EPI_ISL_583823\|2020- | ···················· | ···················· | ···················· | ···················· | ···················· | ···················· | |
| M | 4633 | hCoV-19/Austria/CeMM0979/2020\|EPI_ISL_583824\|2020- | ···················· | ···················· | ···················· | ···················· | ···················· | ···················· | |
| M | 4634 | hCoV-19/Austria/CeMM0980/2020\|EPI_ISL_583825\|2020- | ···················· | ···················· | ···················· | ···················· | ···················· | ···················· | |
| M | 4635 | hCoV-19/Austria/CeMM0981/2020\|EPI_ISL_583826\|2020- | ···················· | ···················· | ···················· | ···················· | ···················· | ···················· | |
| M | 4636 | hCoV-19/Austria/CeMM0982/2020\|EPI_ISL_583827\|2020- | ···················· | ···················· | ···················· | ···················· | ···················· | ···················· | |
| M | 4637 | hCoV-19/Austria/CeMM0984/2020\|EPI_ISL_583828\|2020- | ···················· | ···················· | ···················· | ···················· | ···················· | ···················· | |
| M | 4638 | hCoV-19/Austria/CeMM0985/2020\|EPI_ISL_583829\|2020- | ···················· | ···················· | ···················· | ···················· | ···················· | ···················· | |
| M | 4639 | hCoV-19/Austria/CeMM1096/2020\|EPI_ISL_583892\|2020- | ···················· | ···················· | ···················· | ···················· | ···················· | ···················· | |
| M | 4640 | hCoV-19/Wales/PHWC-16F87F/2020\|EPI_ISL_586029\|2020 | ···················· | ···················· | ···················· | ···················· | ···················· | ···················· | |
| M | 4641 | hCoV-19/Wales/PHWC-16F88E/2020\|EPI_ISL_586030\|2020 | ···················· | ···················· | ···················· | ···················· | ···················· | ···················· | |
| M | 4646 | hCoV-19/Wales/PHWC-16F903/2020\|EPI_ISL_586038\|2020 | ···················· | ···················· | ···················· | ···················· | ···················· | ···················· | |
| M | 4647 | hCoV-19/Wales/PHWC-47CD13/2020\|EPI_ISL_586039\|2020 | ···················· | ···················· | ···················· | ···················· | ···················· | ···················· | |
| M | 4649 | hCoV-19/Wales/PHWC-47CD31/2020\|EPI_ISL_586041\|2020 | ···················· | ···················· | ···················· | ···················· | NNN················· | ···················· | 1 |
| M | 4650 | hCoV-19/Wales/PHWC-47CD6E/2020\|EPI_ISL_586044\|2020 | ···················· | ···················· | ···················· | ···················· | ···················· | ···················· | |
| M | 4651 | hCoV-19/Wales/PHWC-47CD7D/2020\|EPI_ISL_586045\|2020 | ···················· | ···················· | ···················· | ···················· | ···················· | ···················· | |
| M | 4652 | hCoV-19/Wales/PHWC-47CD8C/2020\|EPI_ISL_586046\|2020 | ···················· | ···················· | ···················· | ···················· | ···················· | ···················· | |
| M | 4654 | hCoV-19/Wales/PHWC-47CDC8/2020\|EPI_ISL_586049\|2020 | ···················· | ···················· | ···················· | ···················· | ···················· | ···················· | |
| M | 4655 | hCoV-19/Wales/PHWC-47CDD7/2020\|EPI_ISL_586050\|2020 | ···················· | ···················· | ···················· | ···················· | ···················· | ···················· | |
| M | 4656 | hCoV-19/Wales/PHWC-47CDE6/2020\|EPI_ISL_586051\|2020 | ···················· | ···················· | ···················· | ···················· | ···················· | ···················· | |
| M | 4660 | hCoV-19/Wales/PHWC-47CE5C/2020\|EPI_ISL_586058\|2020 | ···················· | ···················· | ···················· | ···················· | ···················· | ···················· | |
| M | 4661 | hCoV-19/Wales/PHWC-47CE7A/2020\|EPI_ISL_586060\|2020 | ···················· | ···················· | ···················· | ···················· | ···················· | ···················· | |
| M | 4662 | hCoV-19/Wales/PHWC-47CE89/2020\|EPI_ISL_586061\|2020 | ···················· | ···················· | ···················· | ···················· | ···················· | ···················· | |
| M | 4664 | hCoV-19/Wales/PHWC-47CEB6/2020\|EPI_ISL_586064\|2020 | ···················· | ···················· | ···················· | ···················· | ···················· | ···················· | |
| M | 4665 | hCoV-19/Wales/PHWC-47CED4/2020\|EPI_ISL_586066\|2020 | ···················· | ···················· | ···················· | ···················· | ···················· | ···················· | |
| M | 4666 | hCoV-19/Wales/PHWC-47CEF2/2020\|EPI_ISL_586068\|2020 | ···················· | ···················· | ···················· | ···················· | ···················· | ···················· | |
| M | 4667 | hCoV-19/Wales/PHWC-47CF0E/2020\|EPI_ISL_586069\|2020 | ···················· | ···················· | ···················· | ···················· | ···················· | ···················· | |
| M | 4668 | hCoV-19/Wales/PHWC-47CF1D/2020\|EPI_ISL_586070\|2020 | ···················· | ···················· | ···················· | ···················· | ···················· | ···················· | |
| M | 4669 | hCoV-19/Wales/PHWC-47CF2C/2020\|EPI_ISL_586071\|2020 | ···················· | ···················· | ···················· | ···················· | ···················· | ···················· | |
| M | 4670 | hCoV-19/Wales/PHWC-47CF3B/2020\|EPI_ISL_586072\|2020 | ···················· | ···················· | ···················· | ···················· | ···················· | ···················· | |
| M | 4671 | hCoV-19/Wales/PHWC-47CF4A/2020\|EPI_ISL_586073\|2020 | ···················· | ···················· | ···················· | ···················· | ···················· | ···················· | |
| M | 4673 | hCoV-19/Wales/PHWC-47CF68/2020\|EPI_ISL_586075\|2020 | ···················· | ···················· | ···················· | ···················· | ···················· | ···················· | |
| M | 4674 | hCoV-19/Wales/PHWC-47CF77/2020\|EPI_ISL_586076\|2020 | ···················· | ···················· | ···················· | ···················· | ···················· | ···················· | |
| M | 4675 | hCoV-19/Wales/PHWC-47CF86/2020\|EPI_ISL_586077\|2020 | ···················· | ···················· | ···················· | ···················· | ···················· | ···················· | |
| M | 4676 | hCoV-19/Wales/PHWC-47CF95/2020\|EPI_ISL_586078\|2020 | ···················· | ···················· | ···················· | ···················· | ···················· | ···················· | |
| M | 4677 | hCoV-19/Wales/PHWC-47CFB3/2020\|EPI_ISL_586080\|2020 | ···················· | ···················· | ···················· | ···················· | ···················· | ···················· | |
| M | 4679 | hCoV-19/Wales/PHWC-47CFE0/2020\|EPI_ISL_586083\|2020 | ···················· | ···················· | ···················· | ···················· | ···················· | ···················· | |
| M | 4680 | hCoV-19/Wales/PHWC-47CFFF/2020\|EPI_ISL_586084\|2020 | ···················· | ···················· | ···················· | ···················· | ···················· | ···················· | |
| M | 4681 | hCoV-19/Wales/PHWC-47D028/2020\|EPI_ISL_586087\|2020 | ···················· | ···················· | ···················· | ···················· | ···················· | ···················· | |
| M | 4682 | hCoV-19/Wales/PHWC-47D037/2020\|EPI_ISL_586088\|2020 | ···················· | ···················· | ···················· | ···················· | ···················· | ···················· | |
| M | 4684 | hCoV-19/Wales/PHWC-47D091/2020\|EPI_ISL_586093\|2020 | ···················· | ···················· | ···················· | ···················· | ···················· | ···················· | |
| M | 4688 | hCoV-19/Wales/PHWC-47D0EC/2020\|EPI_ISL_586098\|2020 | ···················· | ···················· | ···················· | ···················· | ···················· | ···················· | |
| M | 4689 | hCoV-19/Wales/PHWC-47D0FB/2020\|EPI_ISL_586099\|2020 | ···················· | ···················· | ···················· | ···················· | ···················· | ···················· | |
| M | 4690 | hCoV-19/Wales/PHWC-47D107/2020\|EPI_ISL_586100\|2020 | ···················· | ···················· | ···················· | ···················· | ···················· | ···················· | |
| M | 4693 | hCoV-19/Wales/PHWC-47D2F5/2020\|EPI_ISL_586126\|2020 | ···················· | ···················· | ···················· | ···················· | ···················· | ···················· | |
| M | 4695 | hCoV-19/Wales/PHWC-47D32F/2020\|EPI_ISL_586129\|2020 | ···················· | ···················· | ···················· | ···················· | ···················· | ···················· | |
| M | 4696 | hCoV-19/Wales/PHWC-47D35C/2020\|EPI_ISL_586131\|2020 | ···················· | ···················· | ···················· | ···················· | ···················· | ···················· | |
| M | 4697 | hCoV-19/Wales/PHWC-47D37A/2020\|EPI_ISL_586133\|2020 | ···················· | ···················· | ···················· | ···················· | ···················· | ···················· | |
| M | 4698 | hCoV-19/Wales/PHWC-47D389/2020\|EPI_ISL_586134\|2020 | ···················· | ···················· | ···················· | ···················· | ···················· | ···················· | |
| M | 4700 | hCoV-19/Wales/PHWC-47D3E3/2020\|EPI_ISL_586138\|2020 | ···················· | ···················· | ···················· | ···················· | ···················· | ···················· | |
| M | 4702 | hCoV-19/Wales/PHWC-47D41D/2020\|EPI_ISL_586140\|2020 | ···················· | ···················· | ···················· | ···················· | ···················· | ···················· | |
| M | 4703 | hCoV-19/Wales/PHWC-47D42C/2020\|EPI_ISL_586141\|2020 | ···················· | ···················· | ···················· | ···················· | ···················· | ···················· | |

| | | | | | | | | |
|---|---|---|---|---|---|---|---|---|
| M | 4705 | hCoV-19/Wales/PHWC-47D44A/2020\|EPI_ISL_586143\|2020 | ···················· | ···················· | ····················· | ···················· | ···················· | ···················· |
| M | 4706 | hCoV-19/Wales/PHWC-47D459/2020\|EPI_ISL_586144\|2020 | ···················· | ···················· | ····················· | ···················· | ···················· | ···················· |
| M | 4707 | hCoV-19/Wales/PHWC-47D468/2020\|EPI_ISL_586145\|2020 | ···················· | ···················· | ····················· | ···················· | ···················· | ···················· |
| M | 4708 | hCoV-19/Wales/PHWC-47D477/2020\|EPI_ISL_586146\|2020 | ···················· | ···················· | ····················· | ···················· | ···················· | ···················· |
| M | 4709 | hCoV-19/England/QEUH-A1E102/2020\|EPI_ISL_587358\|20 | ···················· | ···················· | ····················· | ···················· | ···················· | ···················· |
| M | 4710 | hCoV-19/England/QEUH-A1E964/2020\|EPI_ISL_587359\|20 | ···················· | ···················· | ····················· | ···················· | ···················· | ···················· |
| M | 4711 | hCoV-19/England/QEUH-A2188F/2020\|EPI_ISL_587360\|20 | ···················· | ···················· | ····················· | ···················· | ···················· | ···················· |
| M | 4712 | hCoV-19/England/QEUH-A1EDE0/2020\|EPI_ISL_587361\|20 | ···················· | ···················· | ····················· | ···················· | ···················· | ···················· |
| M | 4713 | hCoV-19/England/QEUH-A1DDF0/2020\|EPI_ISL_587362\|20 | ···················· | ···················· | ····················· | ···················· | ···················· | ···················· |
| M | 4714 | hCoV-19/England/QEUH-A1DEB1/2020\|EPI_ISL_587364\|20 | ···················· | ···················· | ····················· | ···················· | ···················· | ···················· |
| M | 4715 | hCoV-19/England/QEUH-A1E8D0/2020\|EPI_ISL_587365\|20 | ···················· | ···················· | ····················· | ···················· | ···················· | ···················· |
| M | 4716 | hCoV-19/England/QEUH-A223D8/2020\|EPI_ISL_587366\|20 | ···················· | ···················· | ····················· | ···················· | ···················· | ···················· |
| M | 4717 | hCoV-19/England/QEUH-A1E287/2020\|EPI_ISL_587367\|20 | ···················· | ···················· | ····················· | ···················· | ···················· | ···················· |
| M | 4718 | hCoV-19/England/QEUH-A1E8B2/2020\|EPI_ISL_587368\|20 | ···················· | ···················· | ····················· | ···················· | ···················· | ···················· |
| M | 4719 | hCoV-19/England/QEUH-A1EB9B/2020\|EPI_ISL_587369\|20 | ···················· | ···················· | ····················· | ···················· | ···················· | ···················· |
| M | 4720 | hCoV-19/England/QEUH-A1EBC8/2020\|EPI_ISL_587370\|20 | ···················· | ···················· | ····················· | ···················· | ···················· | ···················· |
| M | 4721 | hCoV-19/England/QEUH-A1E2A5/2020\|EPI_ISL_587371\|20 | ···················· | ···················· | ····················· | ···················· | ···················· | ···················· |
| M | 4722 | hCoV-19/England/QEUH-A1E74C/2020\|EPI_ISL_587372\|20 | ···················· | ···················· | ····················· | ···················· | ···················· | ···················· |
| M | 4723 | hCoV-19/England/QEUH-A1DE39/2020\|EPI_ISL_587373\|20 | ···················· | ···················· | ····················· | ···················· | ···················· | ···················· |
| M | 4724 | hCoV-19/England/QEUH-A1DB41/2020\|EPI_ISL_587374\|20 | ···················· | ···················· | ····················· | ···················· | ···················· | ···················· |
| M | 4725 | hCoV-19/England/QEUH-A1E919/2020\|EPI_ISL_587375\|20 | ···················· | ···················· | ····················· | ···················· | ···················· | ···················· |
| M | 4726 | hCoV-19/England/QEUH-A21913/2020\|EPI_ISL_587376\|20 | ···················· | ···················· | ····················· | ···················· | ···················· | ···················· |
| M | 4727 | hCoV-19/England/QEUH-A1DAAE/2020\|EPI_ISL_587377\|20 | ···················· | ···················· | ····················· | ···················· | ···················· | ···················· |
| M | 4728 | hCoV-19/England/QEUH-A1EB7D/2020\|EPI_ISL_587378\|20 | ···················· | ···················· | ····················· | ···················· | ···················· | ···················· |
| M | 4729 | hCoV-19/England/QEUH-A1E991/2020\|EPI_ISL_587379\|20 | ···················· | ···················· | ····················· | ···················· | ···T················ | ···················· |
| M | 4730 | hCoV-19/England/QEUH-A1E982/2020\|EPI_ISL_587381\|20 | ···················· | ···················· | ····················· | ···················· | ···················· | ···················· |
| M | 4731 | hCoV-19/England/QEUH-A1DA9F/2020\|EPI_ISL_587384\|20 | ···················· | ···················· | ····················· | ···················· | ···················· | ···················· |
| M | 4732 | hCoV-19/England/QEUH-A1DE57/2020\|EPI_ISL_587385\|20 | ···················· | ···················· | ····················· | ···················· | ···················· | ···················· |
| M | 4733 | hCoV-19/England/QEUH-A1DC02/2020\|EPI_ISL_587387\|20 | ···················· | ···················· | ····················· | ···················· | ···················· | ···················· |
| M | 4734 | hCoV-19/England/QEUH-A1E8FE/2020\|EPI_ISL_587388\|20 | ···················· | ···················· | ····················· | ···················· | ···················· | ···················· |
| M | 4735 | hCoV-19/England/QEUH-A211B1/2020\|EPI_ISL_587389\|20 | ···················· | ···················· | ····················· | ···················· | ···················· | ···················· |
| M | 4736 | hCoV-19/England/QEUH-A22341/2020\|EPI_ISL_587391\|20 | ···················· | ···················· | ····················· | ···················· | ···················· | ···················· |
| M | 4737 | hCoV-19/England/QEUH-A1E023/2020\|EPI_ISL_587392\|20 | ···················· | ···················· | ····················· | ···················· | ···················· | ···················· |
| M | 4738 | hCoV-19/England/QEUH-A1DA71/2020\|EPI_ISL_587393\|20 | ···················· | ···················· | ····················· | ···················· | ···················· | ···················· |
| M | 4739 | hCoV-19/England/QEUH-A1E59D/2020\|EPI_ISL_587394\|20 | ···················· | ···················· | ····················· | ···················· | ···················· | ···················· |
| M | 4740 | hCoV-19/England/QEUH-A21333/2020\|EPI_ISL_587395\|20 | ···················· | ···················· | ····················· | ···················· | ···················· | ···················· |
| M | 4741 | hCoV-19/England/QEUH-A1F462/2020\|EPI_ISL_587396\|20 | ···················· | ···················· | ····················· | ···················· | ···················· | ···················· |
| M | 4742 | hCoV-19/England/QEUH-A2139D/2020\|EPI_ISL_587397\|20 | ···················· | ···················· | ····················· | ···················· | ···················· | ···················· |
| M | 4743 | hCoV-19/England/QEUH-A1DF63/2020\|EPI_ISL_587398\|20 | ···················· | ···················· | ····················· | ···················· | ···················· | ···················· |
| M | 4744 | hCoV-19/England/QEUH-A21F7B/2020\|EPI_ISL_587399\|20 | ···················· | ···················· | ····················· | ···················· | ···················· | ···················· |
| M | 4745 | hCoV-19/England/QEUH-A1DB14/2020\|EPI_ISL_587400\|20 | ···················· | ···················· | ····················· | ···················· | ···················· | ···················· |
| M | 4746 | hCoV-19/England/QEUH-A21597/2020\|EPI_ISL_587402\|20 | ···················· | ···················· | ····················· | ···················· | ···················· | ···················· |
| M | 4747 | hCoV-19/England/QEUH-A2239C/2020\|EPI_ISL_587403\|20 | ···················· | ···················· | ····················· | ···················· | ···················· | ···················· |
| M | 4748 | hCoV-19/England/QEUH-A21834/2020\|EPI_ISL_587404\|20 | ···················· | ···················· | ····················· | ···················· | ···················· | ···················· |
| M | 4749 | hCoV-19/England/QEUH-A212EB/2020\|EPI_ISL_587405\|20 | ···················· | ···················· | ····················· | ···················· | ···················· | ···················· |
| M | 4750 | hCoV-19/England/QEUH-A1E357/2020\|EPI_ISL_587406\|20 | ···················· | ···················· | ····················· | ···················· | ···················· | ···················· |
| M | 4751 | hCoV-19/England/QEUH-A1E928/2020\|EPI_ISL_587407\|20 | ···················· | ···················· | ····················· | ···················· | ···················· | ···················· |
| M | 4752 | hCoV-19/England/QEUH-A1DA62/2020\|EPI_ISL_587408\|20 | ···················· | ···················· | ····················· | ···················· | ···················· | ···················· |
| M | 4753 | hCoV-19/England/QEUH-A1DACC/2020\|EPI_ISL_587409\|20 | ···················· | ···················· | ····················· | ···················· | ···················· | ···················· |
| M | 4754 | hCoV-19/England/QEUH-A2238D/2020\|EPI_ISL_587411\|20 | ···················· | ···················· | ····················· | ···················· | ···················· | ···················· |
| M | 4755 | hCoV-19/England/QEUH-A1DE84/2020\|EPI_ISL_587413\|20 | ···················· | ···················· | ····················· | ···················· | ···················· | ···················· |
| M | 4756 | hCoV-19/England/QEUH-A1E72E/2020\|EPI_ISL_587414\|20 | ···················· | ·A·················· | ····················· | ···················· | ···················· | ··A················· |
| M | 4757 | hCoV-19/England/QEUH-A218DA/2020\|EPI_ISL_587415\|20 | ···················· | ···················· | ····················· | ···················· | ···················· | ···················· |
| M | 4758 | hCoV-19/England/QEUH-A216C1/2020\|EPI_ISL_587416\|20 | ···················· | ···················· | ····················· | ···················· | ···················· | ···················· |
| M | 4759 | hCoV-19/England/QEUH-A1E005/2020\|EPI_ISL_587417\|20 | ···················· | ···················· | ····················· | ···················· | ···················· | ···················· |
| M | 4760 | hCoV-19/England/QEUH-A1EE29/2020\|EPI_ISL_587419\|20 | ···················· | ···················· | ····················· | ···················· | ···················· | ···················· |
| M | 4761 | hCoV-19/England/QEUH-A1E6D6/2020\|EPI_ISL_587420\|20 | ···················· | ···················· | ····················· | ···················· | ···················· | ···················· |
| M | 4762 | hCoV-19/England/QEUH-A21F99/2020\|EPI_ISL_587421\|20 | ···················· | ···················· | ····················· | ···················· | ···················· | ···················· |
| M | 4763 | hCoV-19/England/QEUH-A1DFBE/2020\|EPI_ISL_587422\|20 | ···················· | ···················· | ····················· | ···················· | ···················· | ···················· |
| M | 4764 | hCoV-19/England/QEUH-A218AD/2020\|EPI_ISL_587423\|20 | ···················· | ···················· | ····················· | ···················· | ···················· | ···················· |
| M | 4765 | hCoV-19/England/QEUH-A1DDD2/2020\|EPI_ISL_587424\|20 | ···················· | ···················· | ····················· | ···················· | ···················· | ···················· |
| M | 4766 | hCoV-19/England/QEUH-A21685/2020\|EPI_ISL_587425\|20 | ···················· | ···················· | ····················· | ···················· | ···················· | ···················· |
| M | 4767 | hCoV-19/England/QEUH-A22350/2020\|EPI_ISL_587427\|20 | ···················· | ···················· | ····················· | ···················· | ···················· | ···················· |
| M | 4768 | hCoV-19/England/QEUH-A2237E/2020\|EPI_ISL_587428\|20 | ···················· | ···················· | ····················· | ···················· | ···················· | ···················· |
| M | 4769 | hCoV-19/England/QEUH-A1E384/2020\|EPI_ISL_587430\|20 | ···················· | ···················· | ····················· | ···················· | ···················· | ···················· |
| M | 4770 | hCoV-19/England/QEUH-A1E876/2020\|EPI_ISL_587431\|20 | ···················· | ···················· | ····················· | ···················· | ···················· | ···················· |
| M | 4771 | hCoV-19/England/QEUH-A1E1B7/2020\|EPI_ISL_587432\|20 | ···················· | ···················· | ····················· | ···················· | ···T················ | ···················· |
| M | 4772 | hCoV-19/England/QEUH-A2189E/2020\|EPI_ISL_587433\|20 | ···················· | ···················· | ····················· | ···················· | ···T················ | ···················· |
| M | 4773 | hCoV-19/England/QEUH-A1E24B/2020\|EPI_ISL_587434\|20 | ···················· | ···················· | ····················· | ···················· | ···················· | ···················· |
| M | 4774 | hCoV-19/England/QEUH-A1ED68/2020\|EPI_ISL_587437\|20 | ···················· | ···················· | ····················· | ···················· | ···················· | ···················· |
| M | 4775 | hCoV-19/England/QEUH-A1EDFF/2020\|EPI_ISL_587458\|20 | ···················· | ···················· | ····················· | ···················· | ···················· | ···················· |
| M | 4776 | hCoV-19/England/QEUH-A2236F/2020\|EPI_ISL_587459\|20 | ···················· | ···················· | ····················· | ···················· | ···················· | ···················· |
| M | 4777 | hCoV-19/England/QEUH-A1DE48/2020\|EPI_ISL_587460\|20 | ···················· | ···················· | ····················· | ···················· | ···················· | ···················· |
| M | 4778 | hCoV-19/England/QEUH-A1E4BE/2020\|EPI_ISL_587462\|20 | ···················· | ·A·················· | ····················· | ···················· | ···················· | ··A················· |
| M | 4779 | hCoV-19/England/QEUH-A1DAF9/2020\|EPI_ISL_587463\|20 | ···················· | ···················· | ····················· | ···················· | ···T················ | ···················· |
| M | 4780 | hCoV-19/England/QEUH-A1E3A2/2020\|EPI_ISL_587465\|20 | ···················· | ···················· | ····················· | ···················· | ···················· | ···················· |
| M | 4781 | hCoV-19/England/QEUH-A1DEC0/2020\|EPI_ISL_587467\|20 | ···················· | ···················· | ····················· | ···················· | ···················· | ···················· |
| M | 4782 | hCoV-19/England/QEUH-A2198C/2020\|EPI_ISL_587468\|20 | ···················· | ···················· | ····················· | ···················· | ···················· | ···················· |
| M | 4783 | hCoV-19/England/QEUH-A1F4AE/2020\|EPI_ISL_587469\|20 | ···················· | ···················· | ····················· | ···················· | ···················· | ···················· |
| M | 4784 | hCoV-19/England/QEUH-A213BB/2020\|EPI_ISL_587470\|20 | ···················· | ···················· | ····················· | ···················· | ···················· | ···················· |

| | | | | | | | | | |
|---|---|---|---|---|---|---|---|---|---|
| M | 4785 | hCoV-19/England/QEUH-A1EFDB/2020\|EPI_ISL_587471\|20 | ···················· | ···················· | ···················· | ···················· | ···················· | ···················· | |
| M | 4786 | hCoV-19/England/QEUH-A1DF18/2020\|EPI_ISL_587473\|20 | ···················· | ···················· | ···················· | ···················· | ···················· | ···················· | |
| M | 4787 | hCoV-19/England/QEUH-A1EB8C/2020\|EPI_ISL_587474\|20 | ···················· | ···················· | ···················· | ···················· | ···················· | ···················· | |
| M | 4788 | hCoV-19/England/QEUH-A1E2F0/2020\|EPI_ISL_587475\|20 | ···················· | ···················· | ···················· | ···················· | ···················· | ···················· | |
| M | 4789 | hCoV-19/England/QEUH-A1EFBD/2020\|EPI_ISL_587476\|20 | ···················· | ···················· | ···················· | ···················· | ···················· | ···················· | |
| M | 4790 | hCoV-19/England/QEUH-A1E57F/2020\|EPI_ISL_587478\|20 | ···················· | ···················· | ···················· | ···················· | ···················· | ···················· | |
| M | 4791 | hCoV-19/England/QEUH-9F2F29/2020\|EPI_ISL_588689\|20 | ···················· | ···················· | ···················· | ···················· | ···················· | ···················· | |
| M | 4792 | hCoV-19/England/QEUH-9F384B/2020\|EPI_ISL_588690\|20 | ···················· | ·A·················· | ···················· | ···················· | ···················· | ···A················ | |
| M | 4793 | hCoV-19/England/QEUH-9F54FA/2020\|EPI_ISL_588691\|20 | ···················· | ···················· | ···················· | ···················· | ···················· | ···················· | |
| M | 4794 | hCoV-19/England/QEUH-9F2E68/2020\|EPI_ISL_588692\|20 | ···················· | ···················· | ···················· | ···················· | ···················· | ···················· | |
| M | 4795 | hCoV-19/England/QEUH-9F3E67/2020\|EPI_ISL_588693\|20 | ···················· | ···················· | ···················· | ···················· | ···················· | ···················· | |
| M | 4796 | hCoV-19/England/QEUH-9F31B9/2020\|EPI_ISL_588694\|20 | ···················· | ···················· | ···················· | ···················· | ···················· | ···················· | |
| M | 4797 | hCoV-19/England/QEUH-9F391B/2020\|EPI_ISL_588696\|20 | ···················· | ···················· | ···················· | ···················· | ···················· | ···················· | |
| M | 4798 | hCoV-19/England/QEUH-9F40BB/2020\|EPI_ISL_588698\|20 | ···················· | ·A·················· | ···················· | ···················· | ···················· | ···················· | |
| M | 4799 | hCoV-19/England/QEUH-9F55BB/2020\|EPI_ISL_588699\|20 | ···················· | ···················· | ···················· | ···················· | ···················· | ···················· | |
| M | 4800 | hCoV-19/England/QEUH-9F3D88/2020\|EPI_ISL_588722\|20 | ···················· | ···················· | ···················· | ···················· | ····T··············· | ···················· | |
| M | 4801 | hCoV-19/England/QEUH-9F3C7C/2020\|EPI_ISL_588723\|20 | ···················· | ···················· | ···················· | ···················· | ···················· | ···················· | |
| M | 4802 | hCoV-19/England/QEUH-9F81F0/2020\|EPI_ISL_588725\|20 | ···················· | ···················· | ···················· | ···················· | ···················· | ···················· | |
| M | 4803 | hCoV-19/England/QEUH-9F87FE/2020\|EPI_ISL_588726\|20 | ···················· | ···················· | ···················· | ···················· | ···················· | ···················· | |
| M | 4804 | hCoV-19/England/QEUH-9F7C87/2020\|EPI_ISL_588727\|20 | ···················· | ···················· | ···················· | ···················· | ···················· | ···················· | |
| M | 4805 | hCoV-19/England/QEUH-9F8A04/2020\|EPI_ISL_588730\|20 | ···················· | ···················· | ···················· | ···················· | ···················· | ···················· | |
| M | 4806 | hCoV-19/England/QEUH-9F63ED/2020\|EPI_ISL_588731\|20 | ···················· | ···················· | ···················· | ···················· | ···················· | ···················· | |
| M | 4807 | hCoV-19/England/QEUH-9F82B1/2020\|EPI_ISL_588732\|20 | ···················· | ···················· | ···················· | ···················· | ···················· | ···················· | |
| M | 4808 | hCoV-19/England/QEUH-9F88EC/2020\|EPI_ISL_588733\|20 | ···················· | ···················· | ···················· | ···················· | ···················· | ···················· | |
| M | 4809 | hCoV-19/England/QEUH-9F9098/2020\|EPI_ISL_588734\|20 | ···················· | ···················· | ···················· | ···················· | ···················· | ···················· | |
| M | 4810 | hCoV-19/England/QEUH-9F8CC2/2020\|EPI_ISL_588735\|20 | ···················· | ···················· | ···················· | ···················· | ···················· | ···················· | |
| M | 4811 | hCoV-19/England/QEUH-9F89DA/2020\|EPI_ISL_588736\|20 | ···················· | ···················· | ···················· | ···················· | ···················· | ···················· | |
| M | 4812 | hCoV-19/England/QEUH-9F904D/2020\|EPI_ISL_588737\|20 | ···················· | ···················· | ···················· | ···················· | ···················· | ···················· | |
| M | 4813 | hCoV-19/England/QEUH-9F88FB/2020\|EPI_ISL_588738\|20 | ···················· | ···················· | ···················· | ···················· | ···················· | ···················· | |
| M | 4814 | hCoV-19/England/QEUH-9F871C/2020\|EPI_ISL_588739\|20 | ···················· | ······C············· | ···················· | ···················· | ···················· | ···················· | |
| M | 4815 | hCoV-19/England/QEUH-9F57B5/2020\|EPI_ISL_588740\|20 | ···················· | ·A·················· | ···················· | ···················· | ···················· | ···················· | |
| M | 4816 | hCoV-19/England/QEUH-9F5EDE/2020\|EPI_ISL_588741\|20 | ···················· | ···················· | ···················· | ···················· | ···················· | ···················· | |
| M | 4817 | hCoV-19/England/QEUH-9F9292/2020\|EPI_ISL_588742\|20 | ···················· | ···················· | ···················· | ···················· | ···················· | ···················· | |
| M | 4818 | hCoV-19/England/QEUH-9F89BC/2020\|EPI_ISL_588743\|20 | ···················· | ···················· | ···················· | ···················· | ···················· | ···················· | |
| M | 4819 | hCoV-19/England/QEUH-9F6259/2020\|EPI_ISL_588744\|20 | ···················· | ···················· | ···················· | ···················· | ···················· | ···················· | |
| M | 4820 | hCoV-19/England/QEUH-9F569A/2020\|EPI_ISL_588745\|20 | ···················· | ···················· | ···················· | ···················· | ···················· | ··················Y· | 1 |
| M | 4821 | hCoV-19/England/QEUH-9F8239/2020\|EPI_ISL_588746\|20 | ···················· | ···················· | ···················· | ···················· | ···················· | ···················· | |
| M | 4822 | hCoV-19/England/QEUH-9F8020/2020\|EPI_ISL_588747\|20 | ···················· | ···················· | ···················· | ···················· | ····T··············· | ···················· | |
| M | 4823 | hCoV-19/England/QEUH-9F5612/2020\|EPI_ISL_588748\|20 | ···················· | ···················· | ···················· | ···················· | ···················· | ···················· | |
| M | 4824 | hCoV-19/England/QEUH-9F88BF/2020\|EPI_ISL_588749\|20 | ···················· | ···················· | ···················· | ···················· | ···················· | ···················· | |
| M | 4825 | hCoV-19/England/QEUH-9F609B/2020\|EPI_ISL_588750\|20 | ···················· | ···················· | ···················· | ···················· | ···················· | ···················· | |
| M | 4826 | hCoV-19/England/QEUH-9F6417/2020\|EPI_ISL_588751\|20 | ···················· | ···················· | ···················· | ···················· | ···················· | ···················· | |
| M | 4827 | hCoV-19/England/QEUH-9F5797/2020\|EPI_ISL_588752\|20 | ···················· | ···················· | ···················· | ···················· | ···················· | ···················· | |
| M | 4828 | hCoV-19/England/QEUH-9F7C96/2020\|EPI_ISL_588753\|20 | ···················· | ···················· | ···················· | ···················· | ···················· | ···················· | |
| M | 4829 | hCoV-19/England/QEUH-9F7FBB/2020\|EPI_ISL_588754\|20 | ···················· | ···················· | ···················· | ···················· | ···················· | ···················· | |
| M | 4830 | hCoV-19/England/QEUH-9F8C0E/2020\|EPI_ISL_588755\|20 | ···················· | ···················· | ···················· | ···················· | ···················· | ···················· | |
| M | 4831 | hCoV-19/England/QEUH-9F82DF/2020\|EPI_ISL_588756\|20 | ···················· | ···················· | ···················· | ···················· | ···················· | ···················· | |
| M | 4832 | hCoV-19/England/QEUH-9F8406/2020\|EPI_ISL_588757\|20 | ···················· | ···················· | ···················· | ···················· | ···················· | ···················· | |
| M | 4833 | hCoV-19/England/QEUH-9F8345/2020\|EPI_ISL_588758\|20 | ···················· | ·A·················· | ···················· | ···················· | ···················· | ···················· | |
| M | 4834 | hCoV-19/England/QEUH-9F815A/2020\|EPI_ISL_588759\|20 | ···················· | ···················· | ···················· | ···················· | ···················· | ···················· | |
| M | 4835 | hCoV-19/England/QEUH-9F573D/2020\|EPI_ISL_588760\|20 | ···················· | ···················· | ···················· | ···················· | ···················· | ···················· | |
| M | 4836 | hCoV-19/England/QEUH-9F857C/2020\|EPI_ISL_588761\|20 | ···················· | ···················· | ···················· | ···················· | ···················· | ···················· | |
| M | 4837 | hCoV-19/England/QEUH-9F85C7/2020\|EPI_ISL_588762\|20 | ···················· | ···················· | ···················· | ···················· | ···················· | ···················· | |
| M | 4838 | hCoV-19/England/QEUH-9F5F35/2020\|EPI_ISL_588763\|20 | ···················· | ···················· | ···················· | ···················· | ···················· | ···················· | |
| M | 4839 | hCoV-19/England/QEUH-9F8002/2020\|EPI_ISL_588764\|20 | ···················· | ···················· | ···················· | ···················· | ···················· | ···················· | |
| M | 4840 | hCoV-19/England/QEUH-9F6426/2020\|EPI_ISL_588765\|20 | ···················· | ···················· | ···················· | ···················· | ···················· | ···················· | |
| M | 4841 | hCoV-19/England/QEUH-9F7F9D/2020\|EPI_ISL_588768\|20 | ···················· | ···················· | ···················· | ···················· | ···················· | ···················· | |
| M | 4842 | hCoV-19/England/QEUH-9F898F/2020\|EPI_ISL_588788\|20 | ···················· | ···················· | ···················· | ···················· | ···················· | ···················· | |
| M | 4843 | hCoV-19/England/QEUH-9F9362/2020\|EPI_ISL_588789\|20 | ···················· | ···················· | ···················· | ···················· | ···················· | ···················· | |
| M | 4844 | hCoV-19/England/QEUH-9F8E71/2020\|EPI_ISL_588790\|20 | ···················· | ···················· | ···················· | ···················· | ···················· | ···················· | |
| M | 4845 | hCoV-19/England/QEUH-9F8327/2020\|EPI_ISL_588791\|20 | ···················· | ···················· | ···················· | ···················· | ···················· | ···················· | |
| M | 4846 | hCoV-19/England/QEUH-9F87C1/2020\|EPI_ISL_588793\|20 | ···················· | ···················· | ···················· | ···················· | ···················· | ···················· | |
| M | 4847 | hCoV-19/England/QEUH-9F8846/2020\|EPI_ISL_588794\|20 | ···················· | ···················· | ···················· | ···················· | ···················· | ···················· | |
| M | 4848 | hCoV-19/England/QEUH-9F571F/2020\|EPI_ISL_588795\|20 | ···················· | ···················· | ···················· | ···················· | ···················· | ···················· | |
| M | 4849 | hCoV-19/England/QEUH-9F5F08/2020\|EPI_ISL_588796\|20 | ···················· | ···················· | ···················· | ···················· | ···················· | ···················· | |
| M | 4850 | hCoV-19/England/QEUH-9F6356/2020\|EPI_ISL_588797\|20 | ···················· | ···················· | ···················· | ···················· | ···················· | ···················· | |
| M | 4851 | hCoV-19/England/QEUH-9F8284/2020\|EPI_ISL_588798\|20 | ···················· | ···················· | ···················· | ···················· | ···················· | ···················· | |
| M | 4852 | hCoV-19/England/QEUH-9F6022/2020\|EPI_ISL_588799\|20 | ···················· | ···················· | ···················· | ···················· | ···················· | ···················· | |
| M | 4853 | hCoV-19/Australia/VIC15222/2020\|EPI_ISL_593200\|202 | ···················· | ···················· | ···················· | ···················· | ···················· | ···················· | |
| M | 4854 | hCoV-19/Australia/VIC15231/2020\|EPI_ISL_593207\|202 | ···················· | ···················· | ···················· | ···················· | ···················· | ···················· | |
| M | 4855 | hCoV-19/Australia/VIC15232/2020\|EPI_ISL_593208\|202 | ···················· | ···················· | ···················· | ···················· | ···················· | ···················· | |
| M | 4856 | hCoV-19/Australia/VIC15234/2020\|EPI_ISL_593209\|202 | ···················· | ···················· | ···················· | ···················· | ···················· | ···················· | |
| M | 4857 | hCoV-19/England/BIRM-64F0D/2020\|EPI_ISL_594531\|202 | ···················· | ···················· | ···················· | ···················· | ···················· | ···················· | |
| M | 4858 | hCoV-19/England/BIRM-64FFE/2020\|EPI_ISL_594539\|202 | ···················· | ···················· | ···················· | ···················· | ···················· | ···················· | |
| M | 4859 | hCoV-19/Scotland/QEUH-A01357/2020\|EPI_ISL_595802\|2 | ···················· | ···················· | ···················· | ···················· | ···················· | ···················· | |
| M | 4860 | hCoV-19/England/QEUH-A01366/2020\|EPI_ISL_595803\|20 | ···················· | ···················· | ···················· | ···················· | ···················· | ···················· | |
| M | 4861 | hCoV-19/Scotland/QEUH-A01384/2020\|EPI_ISL_595805\|2 | ···················· | ···················· | ···················· | ···················· | ···················· | ···················· | |
| M | 4862 | hCoV-19/Scotland/QEUH-A013B1/2020\|EPI_ISL_595807\|2 | ···················· | ···················· | ···················· | ···················· | ···················· | ···················· | |
| M | 4863 | hCoV-19/Scotland/QEUH-A013C0/2020\|EPI_ISL_595808\|2 | ···················· | ···················· | ···················· | ···················· | ···················· | ···················· | |
| M | 4864 | hCoV-19/Scotland/QEUH-A013DF/2020\|EPI_ISL_595809\|2 | ···················· | ···················· | ···················· | ···················· | ···················· | ···················· | |

| | | | | | | | |
|---|---|---|---|---|---|---|---|
| M | 4865 hCoV-19/Wales/PHWC-47D18F/2020\|EPI_ISL_586105\|2020 | ···················· | ···················· | ···················· | ···················· | ···················· | ···················· |
| M | 4866 hCoV-19/Wales/PHWC-47D19E/2020\|EPI_ISL_586106\|2020 | ···················· | ···················· | ···················· | ···················· | ···················· | ···················· |
| M | 4867 hCoV-19/Wales/PHWC-47D1AD/2020\|EPI_ISL_586107\|2020 | ···················· | ···················· | ···················· | ···················· | ···················· | ···················· |
| M | 4869 hCoV-19/Wales/PHWC-47D204/2020\|EPI_ISL_586112\|2020 | ···················· | ···················· | ···················· | ···················· | ···················· | ···················· |
| M | 4870 hCoV-19/Wales/PHWC-47D231/2020\|EPI_ISL_586115\|2020 | ···················· | ···················· | ···················· | ···················· | ···················· | ···················· |
| M | 4874 hCoV-19/Wales/PHWC-47D29B/2020\|EPI_ISL_586120\|2020 | ···················· | ···················· | ···················· | ···················· | ···················· | ···················· |
| M | 4876 hCoV-19/Wales/PHWC-47D2D7/2020\|EPI_ISL_586124\|2020 | ···················· | ···················· | ···················· | ···················· | ···················· | ···················· |
| M | 4878 hCoV-19/England/QEUH-A1E199/2020\|EPI_ISL_587435\|20 | ···················· | ···················· | ···················· | ···················· | ···················· | ···················· |
| M | 4879 hCoV-19/England/QEUH-A1E481/2020\|EPI_ISL_587436\|20 | ···················· | ···················· | ···················· | ···················· | ···················· | ···················· |
| M | 4880 hCoV-19/England/QEUH-A1E1D5/2020\|EPI_ISL_587438\|20 | ···················· | ···················· | ···················· | ···················· | ····T··············· | ···················· |
| M | 4881 hCoV-19/England/QEUH-A1EE83/2020\|EPI_ISL_587439\|20 | ···················· | ···················· | ···················· | ···················· | ···················· | ···················· |
| M | 4882 hCoV-19/England/QEUH-A1DEEE/2020\|EPI_ISL_587440\|20 | ···················· | ···················· | ···················· | ···················· | ···················· | ···················· |
| M | 4883 hCoV-19/England/QEUH-A1E454/2020\|EPI_ISL_587442\|20 | ···················· | ···················· | ···················· | ···················· | ···················· | ···················· |
| M | 4884 hCoV-19/England/QEUH-A1DCB7/2020\|EPI_ISL_587443\|20 | ···················· | ···················· | ···················· | ···················· | ···················· | ···················· |
| M | 4885 hCoV-19/England/QEUH-A1E90A/2020\|EPI_ISL_587445\|20 | ···················· | ···················· | ···················· | ···················· | ···················· | ···················· |
| M | 4886 hCoV-19/England/QEUH-A1F4DB/2020\|EPI_ISL_587446\|20 | ···················· | ···················· | ···················· | ···················· | ····T··············· | ···················· |
| M | 4887 hCoV-19/England/QEUH-A1E375/2020\|EPI_ISL_587449\|20 | ···················· | ···················· | ···················· | ···················· | ···················· | ···················· |
| M | 4888 hCoV-19/England/QEUH-A21360/2020\|EPI_ISL_587450\|20 | ···················· | ···················· | ···················· | ···················· | ···················· | ···················· |
| M | 4889 hCoV-19/England/QEUH-A1E867/2020\|EPI_ISL_587452\|20 | ···················· | ·A·················· | ···················· | ···················· | ···················· | ···A················ |
| M | 4890 hCoV-19/England/QEUH-A1DBF6/2020\|EPI_ISL_587453\|20 | ···················· | ···················· | ···················· | ···················· | ···················· | ···················· |
| M | 4891 hCoV-19/England/QEUH-A1DB6F/2020\|EPI_ISL_587454\|20 | ···················· | ···················· | ···················· | ···················· | ···················· | ···················· |
| M | 4892 hCoV-19/England/QEUH-A1E779/2020\|EPI_ISL_587455\|20 | ···················· | ···················· | ···················· | ···················· | ···················· | ···················· |
| M | 4893 hCoV-19/England/QEUH-A1E946/2020\|EPI_ISL_587456\|20 | ···················· | ···················· | ···················· | ···················· | ···················· | ···················· |
| M | 4894 hCoV-19/England/QEUH-9F3F55/2020\|EPI_ISL_588700\|20 | ···················· | ···················· | ···················· | ···················· | ···················· | ···················· |
| M | 4895 hCoV-19/England/QEUH-9F3896/2020\|EPI_ISL_588701\|20 | ···················· | ···················· | ···················· | ···················· | ···················· | ···················· |
| M | 4896 hCoV-19/England/QEUH-9F419A/2020\|EPI_ISL_588702\|20 | ···················· | ···················· | ···················· | ···················· | ···················· | ···················· |
| M | 4897 hCoV-19/England/QEUH-9F3BCA/2020\|EPI_ISL_588703\|20 | ···················· | ···················· | ···················· | ···················· | ···················· | ···················· |
| M | 4898 hCoV-19/England/QEUH-9F3623/2020\|EPI_ISL_588704\|20 | ···················· | ···················· | ···················· | ···················· | ···················· | ···················· |
| M | 4899 hCoV-19/England/QEUH-9F2E4A/2020\|EPI_ISL_588705\|20 | ···················· | ···················· | ···················· | ···················· | ···················· | ···················· |
| M | 4900 hCoV-19/England/QEUH-9F43A3/2020\|EPI_ISL_588706\|20 | ···················· | ···················· | ···················· | ···················· | ···················· | ···················· |
| M | 4901 hCoV-19/England/QEUH-9F3948/2020\|EPI_ISL_588707\|20 | ···················· | ···················· | ···················· | ···················· | ···················· | ···················· |
| M | 4902 hCoV-19/England/QEUH-9F3492/2020\|EPI_ISL_588708\|20 | ···················· | ···················· | ···················· | ···················· | ···················· | ···················· |
| M | 4903 hCoV-19/England/QEUH-9F3386/2020\|EPI_ISL_588709\|20 | ···················· | ···················· | ···················· | ···················· | ···················· | ···················· |
| M | 4904 hCoV-19/England/QEUH-9F327A/2020\|EPI_ISL_588710\|20 | ···················· | ···················· | ···················· | ···················· | ···················· | ···················· |
| M | 4905 hCoV-19/England/QEUH-9F3483/2020\|EPI_ISL_588711\|20 | ···················· | ···················· | ···················· | ···················· | ······T············· | ···················· |
| M | 4906 hCoV-19/England/QEUH-9F41B8/2020\|EPI_ISL_588712\|20 | ···················· | ···················· | ···················· | ···················· | ···················· | ···················· |
| M | 4907 hCoV-19/England/QEUH-9F42E2/2020\|EPI_ISL_588713\|20 | ···················· | ···················· | ···················· | ···················· | ···················· | ···················· |
| M | 4908 hCoV-19/England/QEUH-9F3EEF/2020\|EPI_ISL_588716\|20 | ···················· | ···················· | ···················· | ···················· | ···················· | ···················· |
| M | 4909 hCoV-19/England/QEUH-9F3C6D/2020\|EPI_ISL_588718\|20 | ···················· | ···················· | ···················· | ···················· | ···················· | ···················· |
| M | 4910 hCoV-19/England/QEUH-9F3878/2020\|EPI_ISL_588719\|20 | ···················· | ···················· | ···················· | ···················· | ···················· | ···················· |
| M | 4911 hCoV-19/England/QEUH-9F3AAF/2020\|EPI_ISL_588720\|20 | ···················· | ···················· | ···················· | ···················· | ···················· | ···················· |
| M | 4912 hCoV-19/England/QEUH-9F333B/2020\|EPI_ISL_588721\|20 | ···················· | ···················· | ···················· | ···················· | ···················· | ···················· |
| M | 4913 hCoV-19/England/QEUH-9F5AE9/2020\|EPI_ISL_588766\|20 | ···················· | ···················· | ···················· | ···················· | ···················· | ···················· |
| M | 4914 hCoV-19/England/QEUH-9F5EFC/2020\|EPI_ISL_588767\|20 | ···················· | ···················· | ···················· | ···················· | ···················· | ···················· |
| M | 4915 hCoV-19/England/QEUH-9F613E/2020\|EPI_ISL_588769\|20 | ···················· | ···················· | ···················· | ···················· | ···················· | ···················· |
| M | 4916 hCoV-19/Wales/QEUH-9F5EA1/2020\|EPI_ISL_588770\|2020 | ···················· | ···················· | ···················· | ···················· | ···················· | ···················· |
| M | 4917 hCoV-19/England/QEUH-9F87D0/2020\|EPI_ISL_588771\|20 | ···················· | ···················· | ···················· | ···················· | ···················· | ···················· |
| M | 4918 hCoV-19/England/QEUH-9F7F51/2020\|EPI_ISL_588772\|20 | ···················· | ···················· | ···················· | ···················· | ···················· | ···················· |
| M | 4919 hCoV-19/England/QEUH-9F81B4/2020\|EPI_ISL_588773\|20 | ···················· | ···················· | ···················· | ···················· | ···················· | ···················· |
| M | 4920 hCoV-19/England/QEUH-9F8433/2020\|EPI_ISL_588774\|20 | ···················· | ···················· | ···················· | ···················· | ···················· | ···················· |
| M | 4921 hCoV-19/England/QEUH-9F92ED/2020\|EPI_ISL_588775\|20 | ···················· | ···················· | ···················· | ···················· | ···················· | ···················· |
| M | 4922 hCoV-19/England/QEUH-9F8952/2020\|EPI_ISL_588776\|20 | ···················· | ···················· | ···················· | ···················· | ···················· | ···················· |
| M | 4923 hCoV-19/England/QEUH-9F624A/2020\|EPI_ISL_588777\|20 | ···················· | ···················· | ···················· | ···················· | ···················· | ···················· |
| M | 4924 hCoV-19/England/QEUH-9F8916/2020\|EPI_ISL_588778\|20 | ···················· | ···················· | ···················· | ···················· | ···················· | ···················· |
| M | 4925 hCoV-19/England/QEUH-9F93CC/2020\|EPI_ISL_588779\|20 | ···················· | ···················· | ···················· | ···················· | ···················· | ···················· |
| M | 4926 hCoV-19/England/QEUH-9F90A7/2020\|EPI_ISL_588780\|20 | ···················· | ···················· | ···················· | ···················· | ···················· | ···················· |
| M | 4927 hCoV-19/England/QEUH-9F8934/2020\|EPI_ISL_588781\|20 | ···················· | ·A·················· | ···················· | ···················· | ···················· | ···················· |
| M | 4928 hCoV-19/England/QEUH-9F8318/2020\|EPI_ISL_588782\|20 | ···················· | ···················· | ···················· | ···················· | ···················· | ···················· |
| M | 4929 hCoV-19/England/QEUH-9F880A/2020\|EPI_ISL_588784\|20 | ···················· | ···················· | ···················· | ···················· | ···················· | ···················· |
| M | 4930 hCoV-19/England/QEUH-9F8309/2020\|EPI_ISL_588785\|20 | ···················· | ···················· | ···················· | ···················· | ···················· | ···················· |
| M | 4931 hCoV-19/England/QEUH-9F6295/2020\|EPI_ISL_588786\|20 | ···················· | ···················· | ···················· | ···················· | ···················· | ···················· |
| M | 4932 hCoV-19/England/QEUH-9F623B/2020\|EPI_ISL_588787\|20 | ···················· | ···················· | ···················· | ···················· | ···················· | ···················· |
| M | 4933 hCoV-19/England/MILK-9F25BE/2020\|EPI_ISL_589019\|20 | ···················· | ···················· | ···················· | ···················· | ···················· | ···················· |
| M | 4934 hCoV-19/England/QEUH-9F98EB/2020\|EPI_ISL_589022\|20 | ···················· | ·A·················· | ···················· | ···················· | ···················· | ···················· |
| M | 4935 hCoV-19/England/QEUH-9F96C3/2020\|EPI_ISL_589023\|20 | ···················· | ···················· | ···················· | ···················· | ······T············· | ···················· |
| M | 4936 hCoV-19/England/MILK-9F2220/2020\|EPI_ISL_589024\|20 | ···················· | ···················· | ···················· | ···················· | ···················· | ···················· |
| M | 4937 hCoV-19/England/MILK-9F293A/2020\|EPI_ISL_589026\|20 | ···················· | ···················· | ···················· | ···················· | ···················· | ···················· |
| M | 4938 hCoV-19/England/QEUH-9F7D93/2020\|EPI_ISL_589027\|20 | ···················· | ···················· | ···················· | ···················· | ···················· | ···················· |
| M | 4939 hCoV-19/England/MILK-9F2132/2020\|EPI_ISL_589028\|20 | ···················· | ···················· | ···················· | ···················· | ···················· | ···················· |
| M | 4940 hCoV-19/England/MILK-9F2554/2020\|EPI_ISL_589029\|20 | ···················· | ···················· | ···················· | ···················· | ···················· | ···················· |
| M | 4941 hCoV-19/England/QEUH-9F95D5/2020\|EPI_ISL_589033\|20 | ···················· | ···················· | ···················· | ···················· | ······T············· | ···················· |
| M | 4942 hCoV-19/England/MILK-9F2581/2020\|EPI_ISL_589034\|20 | ···················· | ···················· | ···················· | ···················· | ···················· | ···················· |
| M | 4943 hCoV-19/England/QEUH-9F7AE7/2020\|EPI_ISL_589035\|20 | ···················· | ···················· | ···················· | ···················· | ····T··············· | ···················· |
| M | 4944 hCoV-19/England/QEUH-9F9CA3/2020\|EPI_ISL_589036\|20 | ···················· | ···················· | ···················· | ···················· | ···················· | ···················· |
| M | 4945 hCoV-19/England/MILK-9F21E7/2020\|EPI_ISL_589039\|20 | ···················· | ·A·················· | ···················· | ···················· | ···················· | ···A················ |
| M | 4946 hCoV-19/England/MILK-9F2606/2020\|EPI_ISL_589040\|20 | ···················· | ···················· | ···················· | ···················· | ···················· | ···················· |
| M | 4947 hCoV-19/England/MILK-9F2642/2020\|EPI_ISL_589041\|20 | ···················· | ·A·················· | ···················· | ···················· | ···················· | ···A················ |
| M | 4948 hCoV-19/England/QEUH-9F7935/2020\|EPI_ISL_589042\|20 | ···················· | ···················· | ···················· | ···················· | ···················· | ···················· |
| M | 4949 hCoV-19/England/MILK-9F2A19/2020\|EPI_ISL_589043\|20 | ···················· | ···················· | ···················· | ···················· | ···················· | ···················· |
| M | 4950 hCoV-19/England/QEUH-9F95C6/2020\|EPI_ISL_589044\|20 | ···················· | ···················· | ···················· | ···················· | ···················· | ···················· |

| | | | | | | | | |
|---|---|---|---|---|---|---|---|---|
| M | 4951 | hCoV-19/England/QEUH-9F96D2/2020\|EPI_ISL_589045\|20 | ···················· | ···················· | ····················· | ···················· | ···················· | ···················· |
| M | 4952 | hCoV-19/England/QEUH-9F7BC6/2020\|EPI_ISL_589047\|20 | ···················· | ···················· | ····················· | ···················· | ···················· | ···················· |
| M | 4953 | hCoV-19/England/QEUH-9F72D0/2020\|EPI_ISL_589048\|20 | ···················· | ···················· | ····················· | ···················· | ···················· | ···················· |
| M | 4954 | hCoV-19/England/QEUH-9F7908/2020\|EPI_ISL_589049\|20 | ···················· | ···················· | ····················· | ···················· | ···················· | ···················· |
| M | 4955 | hCoV-19/England/MILK-9F2466/2020\|EPI_ISL_589051\|20 | ···················· | ···················· | ····················· | ···················· | ···················· | ···················· |
| M | 4956 | hCoV-19/England/QEUH-9F7C0F/2020\|EPI_ISL_589053\|20 | ···················· | ···················· | ····················· | ···················· | ···················· | ···················· |
| M | 4957 | hCoV-19/England/QEUH-9F9960/2020\|EPI_ISL_589054\|20 | ···················· | ···················· | ····················· | ···················· | ···················· | ···················· |
| M | 4958 | hCoV-19/England/QEUH-9F9766/2020\|EPI_ISL_589055\|20 | ···················· | ···················· | ····················· | ···················· | ···················· | ···················· |
| M | 4959 | hCoV-19/England/MILK-9F283D/2020\|EPI_ISL_589057\|20 | ···················· | ···················· | ····················· | ···················· | ···················· | ···················· |
| M | 4960 | hCoV-19/England/QEUH-9F9872/2020\|EPI_ISL_589058\|20 | ···················· | ···················· | ····················· | ···················· | ···················· | ···················· |
| M | 4961 | hCoV-19/Wales/QEUH-9CFF8F/2020\|EPI_ISL_590001\|2020 | ···················· | ···················· | ····················· | ···················· | ······T············· | ···················· |
| M | 4962 | hCoV-19/England/QEUH-9D0871/2020\|EPI_ISL_590002\|20 | ···················· | ···················· | ····················· | ···················· | ···················· | ···················· |
| M | 4963 | hCoV-19/England/QEUH-9D039E/2020\|EPI_ISL_590003\|20 | ···················· | ···················· | ····················· | ···················· | ···················· | ···················· |
| M | 4964 | hCoV-19/England/QEUH-9D0282/2020\|EPI_ISL_590004\|20 | ···················· | ···················· | ····················· | ···················· | ···················· | ···················· |
| M | 4965 | hCoV-19/England/QEUH-9D07CF/2020\|EPI_ISL_590005\|20 | ···················· | ·A·················· | ····················· | ···················· | ···················· | ···················· |
| M | 4966 | hCoV-19/England/QEUH-9D049B/2020\|EPI_ISL_590006\|20 | ···················· | ···················· | ····················· | ···················· | ···················· | ···················· |
| M | 4967 | hCoV-19/England/QEUH-9D0738/2020\|EPI_ISL_590007\|20 | ···················· | ···················· | ····················· | ···················· | ···················· | ···················· |
| M | 4968 | hCoV-19/England/QEUH-9D0079/2020\|EPI_ISL_590009\|20 | ···················· | ···················· | ····················· | ···················· | ···················· | ···················· |
| M | 4969 | hCoV-19/England/QEUH-9D112A/2020\|EPI_ISL_590010\|20 | ···················· | ···················· | ····················· | ···················· | ···················· | ···················· |
| M | 4970 | hCoV-19/England/QEUH-9D0D54/2020\|EPI_ISL_590011\|20 | ···················· | ···················· | ····················· | ···················· | ···················· | ···················· |
| M | 4971 | hCoV-19/England/QEUH-9D1148/2020\|EPI_ISL_590012\|20 | ···················· | ···················· | ····················· | ···················· | ····T··············· | ···················· |
| M | 4972 | hCoV-19/England/QEUH-9CFACA/2020\|EPI_ISL_590013\|20 | ···················· | ···················· | ····················· | ···················· | ···················· | ···················· |
| M | 4973 | hCoV-19/England/QEUH-9CE4BE/2020\|EPI_ISL_590016\|20 | ···················· | ···················· | ····················· | ···················· | ···················· | ···················· |
| M | 4974 | hCoV-19/England/QEUH-9CF972/2020\|EPI_ISL_590021\|20 | ···················· | ···················· | ····················· | ···················· | ···················· | ···················· |
| M | 4975 | hCoV-19/England/QEUH-9CF5AB/2020\|EPI_ISL_590023\|20 | ···················· | ···················· | ····················· | ···················· | ···················· | ···················· |
| M | 4976 | hCoV-19/England/QEUH-9CE1F3/2020\|EPI_ISL_590024\|20 | ···················· | ···················· | ····················· | ···················· | ···················· | ···················· |
| M | 4977 | hCoV-19/England/QEUH-9CF3FC/2020\|EPI_ISL_590025\|20 | ···················· | ···················· | ····················· | ···················· | ···················· | ···················· |
| M | 4978 | hCoV-19/England/QEUH-9CE867/2020\|EPI_ISL_590026\|20 | ···················· | ···················· | ····················· | ···················· | ···················· | ···················· |
| M | 4979 | hCoV-19/England/QEUH-9CE75B/2020\|EPI_ISL_590027\|20 | ···················· | ···················· | ····················· | ···················· | ····T··············· | ···················· |
| M | 4980 | hCoV-19/England/QEUH-9CE472/2020\|EPI_ISL_590028\|20 | ···················· | ···················· | ····················· | ···················· | ···················· | ···················· |
| M | 4981 | hCoV-19/England/QEUH-9CF277/2020\|EPI_ISL_590029\|20 | ···················· | ···················· | ····················· | ···················· | ····T··············· | ···················· |
| M | 4982 | hCoV-19/England/QEUH-9CED3B/2020\|EPI_ISL_590030\|20 | ···················· | ·A·················· | ····················· | ···················· | ···················· | ···················· |
| M | 4983 | hCoV-19/England/QEUH-9CEB40/2020\|EPI_ISL_590031\|20 | ···················· | ···················· | ····················· | ···················· | ····T··············· | ···················· |
| M | 4984 | hCoV-19/England/QEUH-9CEE65/2020\|EPI_ISL_590032\|20 | ···················· | ···················· | ····················· | ···················· | ···················· | ···················· |
| M | 4985 | hCoV-19/England/QEUH-9CF0C8/2020\|EPI_ISL_590033\|20 | ···················· | ···················· | ····················· | ···················· | ···················· | ···················· |
| M | 4986 | hCoV-19/England/QEUH-9CEB6E/2020\|EPI_ISL_590034\|20 | ···················· | ···················· | ····················· | ···················· | ···················· | ···················· |
| M | 4987 | hCoV-19/England/QEUH-9CE5AC/2020\|EPI_ISL_590037\|20 | ···················· | ···················· | ····················· | ···················· | ···················· | ···················· |
| M | 4988 | hCoV-19/England/QEUH-9CF7F0/2020\|EPI_ISL_590038\|20 | ···················· | ···················· | ····················· | ···················· | ···················· | ···················· |
| M | 4989 | hCoV-19/England/QEUH-9CE7F1/2020\|EPI_ISL_590039\|20 | ···················· | ···················· | ····················· | ···················· | ···················· | ···················· |
| M | 4990 | hCoV-19/England/QEUH-9CEC2F/2020\|EPI_ISL_590041\|20 | ···················· | ···················· | ····················· | ···················· | ···················· | ···················· |
| M | 4991 | hCoV-19/Italy/TUS-UOS-19/2020\|EPI_ISL_591330\|2020- | ···················· | ···················· | ····················· | ···················· | ···················· | ···················· |
| M | 4992 | hCoV-19/Italy/TUS-UOS-21/2020\|EPI_ISL_591331\|2020- | ···················· | ···················· | ····················· | ···················· | ···················· | ···················· |
| M | 4993 | hCoV-19/Italy/TUS-UOS-4/2020\|EPI_ISL_591332\|2020-0 | ···················· | ···················· | ····················· | ···················· | ···················· | ···················· |
| M | 4994 | hCoV-19/Italy/TUS-UOS-6/2020\|EPI_ISL_591333\|2020-0 | ···················· | ···················· | ····················· | ···················· | ···················· | ···················· |
| M | 4995 | hCoV-19/Italy/TUS-UOS-07/2020\|EPI_ISL_591334\|2020- | ···················· | ···················· | ····················· | ···················· | ···················· | ···················· |
| M | 4996 | hCoV-19/Italy/TUS-UOS-8/2020\|EPI_ISL_591335\|2020-0 | ···················· | NN·················· | ····················· | ···················· | ···················· | ···················· |
| M | 4997 | hCoV-19/Japan/IC-0001/2020\|EPI_ISL_591347\|2020-03 | ···················· | ···················· | ····················· | ···················· | ···················· | ···················· |
| M | 4998 | hCoV-19/Japan/IC-0004/2020\|EPI_ISL_591350\|2020-03 | ···················· | ···················· | ····················· | ···················· | ···················· | ···················· |
| M | 4999 | hCoV-19/Japan/IC-0005/2020\|EPI_ISL_591351\|2020-04 | ···················· | ···················· | ····················· | ···················· | ···················· | ···················· |
| M | 5000 | hCoV-19/Japan/IC-0006/2020\|EPI_ISL_591352\|2020-06 | ···················· | ···················· | ····················· | ···················· | ···················· | ···················· |
| M | 5001 | hCoV-19/Japan/IC-0007/2020\|EPI_ISL_591353\|2020-06 | ···················· | ···················· | ····················· | ···················· | ···················· | ···················· |
| M | 5002 | hCoV-19/Japan/IC-0008/2020\|EPI_ISL_591354\|2020-03 | ···················· | ···················· | ····················· | ···················· | ···················· | ···················· |
| M | 5003 | hCoV-19/Japan/IC-0009/2020\|EPI_ISL_591355\|2020-03 | ···················· | ···················· | ····················· | ···················· | ···················· | ···················· |
| M | 5004 | hCoV-19/Japan/IC-0010/2020\|EPI_ISL_591356\|2020-03 | ···················· | ···················· | ····················· | ···················· | ···················· | ···················· |
| M | 5005 | hCoV-19/Japan/IC-0011/2020\|EPI_ISL_591357\|2020-03 | ···················· | ···················· | ····················· | ···················· | ···················· | ···················· |
| M | 5006 | hCoV-19/Japan/IC-0012/2020\|EPI_ISL_591358\|2020-03 | ···················· | ···················· | ····················· | ···················· | ···················· | ···················· |
| M | 5007 | hCoV-19/Japan/IC-0014/2020\|EPI_ISL_591359\|2020-03 | ···················· | ···················· | ····················· | ···················· | ···················· | ···················· |
| M | 5008 | hCoV-19/Japan/IC-0015/2020\|EPI_ISL_591360\|2020-03 | ···················· | ···················· | ····················· | ···················· | ···················· | ···················· |
| M | 5009 | hCoV-19/Japan/IC-0016/2020\|EPI_ISL_591361\|2020-04 | ···················· | ···················· | ····················· | ···················· | ···················· | ···················· |
| M | 5010 | hCoV-19/Japan/IC-0017/2020\|EPI_ISL_591362\|2020-04 | ···················· | ···················· | ····················· | ···················· | ···················· | ···················· |
| M | 5011 | hCoV-19/Japan/IC-0018/2020\|EPI_ISL_591363\|2020-04 | ···················· | ···················· | ····················· | ···················· | ···················· | ···················· |
| M | 5012 | hCoV-19/Japan/IC-0019/2020\|EPI_ISL_591364\|2020-04 | ···················· | ···················· | ····················· | ···················· | ···················· | ···················· |
| M | 5013 | hCoV-19/Japan/IC-0020/2020\|EPI_ISL_591365\|2020-04 | ···················· | ···················· | ····················· | ···················· | ···················· | ···················· |
| M | 5014 | hCoV-19/Japan/IC-0021/2020\|EPI_ISL_591366\|2020-04 | ···················· | ···················· | ····················· | ···················· | ···················· | ···················· |
| M | 5015 | hCoV-19/Japan/IC-0023/2020\|EPI_ISL_591367\|2020-04 | ···················· | ···················· | ····················· | ···················· | ···················· | ···················· |
| M | 5016 | hCoV-19/Japan/IC-0024/2020\|EPI_ISL_591368\|2020-04 | ···················· | ···················· | ····················· | ···················· | ···················· | ···················· |
| M | 5017 | hCoV-19/Japan/IC-0025/2020\|EPI_ISL_591369\|2020-04 | ···················· | ···················· | ····················· | ···················· | ···················· | ···················· |
| M | 5018 | hCoV-19/Japan/IC-0026/2020\|EPI_ISL_591370\|2020-05 | ···················· | ···················· | ····················· | ···················· | ···················· | ···················· |
| M | 5019 | hCoV-19/Japan/IC-0027/2020\|EPI_ISL_591371\|2020-05 | ···················· | ···················· | ····················· | ···················· | ···················· | ···················· |
| M | 5020 | hCoV-19/Japan/IC-0028/2020\|EPI_ISL_591372\|2020-05 | ···················· | ···················· | ····················· | ···················· | ···················· | ···················· |
| M | 5021 | hCoV-19/Australia/VIC15280/2020\|EPI_ISL_593232\|202 | ···················· | ···················· | ····················· | ···················· | ···················· | ···················· |
| M | 5022 | hCoV-19/Australia/VIC15282/2020\|EPI_ISL_593233\|202 | ···················· | ···················· | ····················· | ···················· | ···················· | ···················· |
| M | 5023 | hCoV-19/Australia/VIC15284/2020\|EPI_ISL_593234\|202 | ···················· | ···················· | ····················· | ···················· | ···················· | ···················· |
| M | 5024 | hCoV-19/Australia/VIC15286/2020\|EPI_ISL_593235\|202 | ···················· | ···················· | ····················· | ···················· | ···················· | ···················· |
| M | 5025 | hCoV-19/Australia/VIC15289/2020\|EPI_ISL_593236\|202 | ···················· | ···················· | ····················· | ···················· | ···················· | ···················· |
| M | 5026 | hCoV-19/Australia/VIC15291/2020\|EPI_ISL_593237\|202 | ···················· | ···················· | ····················· | ···················· | ···················· | ···················· |
| M | 5027 | hCoV-19/Australia/VIC15292/2020\|EPI_ISL_593238\|202 | ···················· | ···················· | ····················· | ···················· | ···················· | ···················· |
| M | 5029 | hCoV-19/Australia/VIC15297/2020\|EPI_ISL_593240\|202 | ···················· | ···················· | ····················· | ···················· | ···················· | ···················· |
| M | 5030 | hCoV-19/Australia/VIC15299/2020\|EPI_ISL_593241\|202 | ···················· | ···················· | ····················· | ···················· | ···················· | ···················· |
| M | 5031 | hCoV-19/Australia/VIC15300/2020\|EPI_ISL_593242\|202 | ···················· | ···················· | ····················· | ···················· | ···················· | ···················· |

| | | | | | | | |
|---|---|---|---|---|---|---|---|
| M | 5032 hCoV-19/Australia/VIC15301/2020\|EPI_ISL_593243\|202 | .................... | .................... | .................... | .................... | .................... | .................... |
| M | 5033 hCoV-19/Australia/VIC15302/2020\|EPI_ISL_593244\|202 | .................... | .................... | .................... | .................... | .................... | .................... |
| M | 5034 hCoV-19/Australia/VIC15305/2020\|EPI_ISL_593245\|202 | .................... | .................... | .................... | .................... | .................... | .................... |
| M | 5035 hCoV-19/Australia/VIC15312/2020\|EPI_ISL_593246\|202 | .................... | .................... | .................... | .................... | .................... | .................... |
| M | 5036 hCoV-19/Australia/VIC15315/2020\|EPI_ISL_593247\|202 | .................... | .................... | .................... | .................... | .................... | .................... |
| M | 5037 hCoV-19/Australia/VIC15327/2020\|EPI_ISL_593248\|202 | .................... | .................... | .................... | .................... | .................... | .................... |
| M | 5038 hCoV-19/Australia/VIC15328/2020\|EPI_ISL_593249\|202 | .................... | .................... | .................... | .................... | .................... | .................... |
| M | 5039 hCoV-19/Australia/VIC15329/2020\|EPI_ISL_593250\|202 | .................... | .................... | .................... | .................... | .................... | .................... |
| M | 5040 hCoV-19/Australia/VIC15330/2020\|EPI_ISL_593251\|202 | .................... | .................... | .................... | .................... | .................... | .................... |
| M | 5041 hCoV-19/Australia/VIC15331/2020\|EPI_ISL_593252\|202 | .................... | .................... | .................... | .................... | .................... | .................... |
| M | 5042 hCoV-19/Australia/VIC15332/2020\|EPI_ISL_593253\|202 | .................... | .................... | .................... | .................... | .................... | .................... |
| M | 5043 hCoV-19/Australia/VIC15333/2020\|EPI_ISL_593254\|202 | .................... | .................... | .................... | .................... | ..Y................. | .................... |
| M | 5044 hCoV-19/Australia/VIC15334/2020\|EPI_ISL_593255\|202 | .................... | .................... | .................... | .................... | .................... | .................... |
| M | 5045 hCoV-19/Australia/VIC15336/2020\|EPI_ISL_593256\|202 | .................... | .................... | .................... | .................... | .................... | .................... |
| M | 5046 hCoV-19/Australia/VIC15353/2020\|EPI_ISL_593257\|202 | .................... | .................... | .................... | .................... | .................... | .................... |
| M | 5047 hCoV-19/Australia/VIC15356/2020\|EPI_ISL_593258\|202 | .................... | .................... | .................... | .................... | .................... | .................... |
| M | 5048 hCoV-19/Australia/VIC15357/2020\|EPI_ISL_593259\|202 | .................... | .................... | .................... | .................... | .................... | .................... |
| M | 5049 hCoV-19/Australia/VIC15360/2020\|EPI_ISL_593260\|202 | .................... | .................... | .................... | .................... | .................... | .................... |
| M | 5050 hCoV-19/Australia/VIC15361/2020\|EPI_ISL_593261\|202 | .................... | .................... | .................... | .................... | .................... | .................... |
| M | 5051 hCoV-19/Australia/VIC15362/2020\|EPI_ISL_593262\|202 | .................... | .................... | .................... | .................... | .................... | .................... |
| M | 5052 hCoV-19/Australia/VIC15363/2020\|EPI_ISL_593263\|202 | .................... | .................... | .................... | .................... | .................... | .................... |
| M | 5053 hCoV-19/Australia/VIC15364/2020\|EPI_ISL_593264\|202 | .................... | .................... | .................... | .................... | .................... | .................... |
| M | 5054 hCoV-19/Australia/VIC15367/2020\|EPI_ISL_593265\|202 | .................... | .................... | .................... | .................... | .................... | .................... |
| M | 5055 hCoV-19/Australia/VIC15369/2020\|EPI_ISL_593266\|202 | .................... | .................... | .................... | .................... | .................... | .................... |
| M | 5056 hCoV-19/Australia/VIC15370/2020\|EPI_ISL_593267\|202 | .................... | .................... | .................... | .................... | .................... | .................... |
| M | 5057 hCoV-19/Australia/VIC15371/2020\|EPI_ISL_593268\|202 | .................... | .................... | .................... | .................... | .................... | .................... |
| M | 5058 hCoV-19/Australia/VIC15372/2020\|EPI_ISL_593269\|202 | .................... | .................... | .................... | .................... | .................... | .................... |
| M | 5059 hCoV-19/Australia/VIC15375/2020\|EPI_ISL_593270\|202 | .................... | .................... | .................... | .................... | .................... | .................... |
| M | 5060 hCoV-19/Australia/VIC15377/2020\|EPI_ISL_593271\|202 | .................... | .................... | .................... | .................... | .................... | .................... |
| M | 5061 hCoV-19/Australia/VIC15378/2020\|EPI_ISL_593272\|202 | .................... | .................... | .................... | .................... | .................... | .................... |
| M | 5062 hCoV-19/Australia/VIC15381/2020\|EPI_ISL_593273\|202 | .................... | .................... | .................... | .................... | .................... | .................... |
| M | 5063 hCoV-19/Australia/VIC15383/2020\|EPI_ISL_593274\|202 | .................... | .................... | .................... | .................... | .................... | .................... |
| M | 5064 hCoV-19/Australia/VIC15384/2020\|EPI_ISL_593275\|202 | .................... | .................... | .................... | .................... | .................... | .................... |
| M | 5065 hCoV-19/Australia/VIC15385/2020\|EPI_ISL_593276\|202 | .................... | .................... | .................... | .................... | .................... | .................... |
| M | 5066 hCoV-19/Australia/VIC15386/2020\|EPI_ISL_593277\|202 | .................... | .................... | .................... | .................... | .................... | .................... |
| M | 5067 hCoV-19/Australia/VIC15387/2020\|EPI_ISL_593278\|202 | .................... | .................... | .................... | .................... | .................... | .................... |
| M | 5068 hCoV-19/Australia/VIC15388/2020\|EPI_ISL_593279\|202 | .................... | .................... | .................... | .................... | .................... | .................... |
| M | 5069 hCoV-19/Australia/VIC15392/2020\|EPI_ISL_593280\|202 | .................... | .................... | .................... | .................... | .................... | .................... |
| M | 5071 hCoV-19/Australia/VIC15394/2020\|EPI_ISL_593282\|202 | .................... | .................... | .................... | .................... | .................... | .................... |
| M | 5072 hCoV-19/Australia/VIC15395/2020\|EPI_ISL_593283\|202 | .................... | .................... | .................... | .................... | .................... | .................... |
| M | 5074 hCoV-19/Australia/VIC15401/2020\|EPI_ISL_593285\|202 | .................... | .................... | .................... | .................... | .................... | .................... |
| M | 5075 hCoV-19/Australia/VIC15402/2020\|EPI_ISL_593286\|202 | .................... | ......A............. | .................... | .................... | .................... | .................... |
| M | 5076 hCoV-19/Australia/VIC15403/2020\|EPI_ISL_593287\|202 | .................... | .................... | .................... | .................... | .................... | .................... |
| M | 5077 hCoV-19/Australia/VIC15404/2020\|EPI_ISL_593288\|202 | .................... | .................... | .................... | .................... | .................... | .................... |
| M | 5078 hCoV-19/Australia/VIC15406/2020\|EPI_ISL_593289\|202 | .................... | .................... | .................... | .................... | .................... | .................... |
| M | 5079 hCoV-19/Australia/VIC15408/2020\|EPI_ISL_593290\|202 | .................... | .................... | .................... | .................... | .................... | .................... |
| M | 5080 hCoV-19/Australia/VIC15410/2020\|EPI_ISL_593291\|202 | .................... | .................... | .................... | .................... | .................... | .................... |
| M | 5081 hCoV-19/Australia/VIC15411/2020\|EPI_ISL_593292\|202 | .................... | .................... | .................... | .................... | .................... | .................... |
| M | 5082 hCoV-19/Australia/VIC15412/2020\|EPI_ISL_593293\|202 | .................... | .................... | .................... | .................... | .................... | .................... |
| M | 5084 hCoV-19/Australia/VIC15414/2020\|EPI_ISL_593295\|202 | .................... | .................... | .................... | .................... | .................... | .................... |
| M | 5085 hCoV-19/Australia/VIC15416/2020\|EPI_ISL_593296\|202 | .................... | .................... | .................... | .................... | .................... | .................... |
| M | 5086 hCoV-19/Australia/VIC15417/2020\|EPI_ISL_593297\|202 | .................... | .................... | .................... | .................... | .................... | .................... |
| M | 5087 hCoV-19/Australia/VIC15418/2020\|EPI_ISL_593298\|202 | .................... | .................... | .................... | .................... | .................... | .................... |
| M | 5088 hCoV-19/Australia/VIC15420/2020\|EPI_ISL_593299\|202 | .................... | .................... | .................... | .................... | .................... | .................... |
| M | 5089 hCoV-19/England/204221492/2020\|EPI_ISL_593991\|2020 | .................... | .................... | .................... | .................... | .................... | .................... |
| M | 5090 hCoV-19/England/204221495/2020\|EPI_ISL_593993\|2020 | .................... | .................... | .................... | .................... | .................... | .................... |
| M | 5091 hCoV-19/England/204221496/2020\|EPI_ISL_593994\|2020 | .................... | .................... | .................... | .................... | .................... | .................... |
| M | 5092 hCoV-19/England/204221497/2020\|EPI_ISL_593995\|2020 | .................... | .................... | .................... | .................... | .................... | .................... |
| M | 5093 hCoV-19/England/204221498/2020\|EPI_ISL_593996\|2020 | .................... | .................... | .................... | .................... | .................... | .................... |
| M | 5094 hCoV-19/England/204221499/2020\|EPI_ISL_593997\|2020 | .................... | .................... | .................... | .................... | .................... | .................... |
| M | 5095 hCoV-19/England/BIRM-65546/2020\|EPI_ISL_594564\|202 | .................... | .................... | .................... | .................... | .................... | .................... |
| M | 5096 hCoV-19/England/BIRM-65573/2020\|EPI_ISL_594566\|202 | .................... | .A.................. | .................... | .................... | .................... | ..A................. |
| M | 5097 hCoV-19/England/BIRM-65591/2020\|EPI_ISL_594568\|202 | .................... | .A.................. | .................... | .................... | .................... | ..A................. |
| M | 5098 hCoV-19/England/BIRM-655CE/2020\|EPI_ISL_594569\|202 | .................... | .A.................. | .................... | .................... | .................... | ..A................. |
| M | 5099 hCoV-19/England/BIRM-655FB/2020\|EPI_ISL_594570\|202 | .................... | .................... | .................... | .................... | .................... | .................... |
| M | 5101 hCoV-19/England/BIRM-65661/2020\|EPI_ISL_594574\|202 | .................... | .A.................. | .................... | .................... | .................... | ..A................. |
| M | 5102 hCoV-19/England/BIRM-6568F/2020\|EPI_ISL_594575\|202 | .................... | .A.................. | .................... | .................... | .................... | ..N................. |
| M | 5104 hCoV-19/England/BIRM-65722/2020\|EPI_ISL_594577\|202 | .................... | .................... | .................... | .................... | .................... | .................... |
| M | 5105 hCoV-19/England/BIRM-65731/2020\|EPI_ISL_594578\|202 | .................... | .................... | .................... | .................... | .................... | .................... |
| M | 5109 hCoV-19/England/BIRM-6584D/2020\|EPI_ISL_594586\|202 | .................... | .A.................. | .................... | .................... | .................... | ..A................. |
| M | 5110 hCoV-19/England/BIRM-6586B/2020\|EPI_ISL_594588\|202 | .................... | .................... | .................... | .................... | .................... | .................... |
| M | 5111 hCoV-19/England/BIRM-6587A/2020\|EPI_ISL_594589\|202 | .................... | .................... | .................... | .................... | .................... | .................... |
| M | 5114 hCoV-19/England/BIRM-6594A/2020\|EPI_ISL_594598\|202 | .................... | .................... | .................... | .................... | .................... | .................... |
| M | 5116 hCoV-19/England/QEUH-A314A6/2020\|EPI_ISL_595894\|20 | .................... | .................... | .................... | .................... | .................... | .................... |
| M | 5117 hCoV-19/Scotland/QEUH-A314C4/2020\|EPI_ISL_595895\|2 | .................... | .................... | .................... | .................... | .................... | .................... |
| M | 5118 hCoV-19/Scotland/QEUH-A314D3/2020\|EPI_ISL_595896\|2 | .................... | .................... | .................... | .................... | .................... | .................... |
| M | 5119 hCoV-19/Scotland/QEUH-A314E2/2020\|EPI_ISL_595897\|2 | .................... | .................... | .................... | .................... | .................... | .................... |
| M | 5120 hCoV-19/Scotland/QEUH-A3152B/2020\|EPI_ISL_595899\|2 | .................... | .................... | .................... | .................... | .................... | .................... |
| M | 5121 hCoV-19/England/ALDP-9E9954/2020\|EPI_ISL_584268\|20 | .................... | .................... | .................... | .................... | .................... | .................... |
| M | 5122 hCoV-19/England/ALDP-9E9963/2020\|EPI_ISL_584269\|20 | .................... | .................... | .................... | .................... | .................... | .................... |

| | | | | | | | |
|---|---|---|---|---|---|---|---|
| M | 5123 hCoV-19/England/ALDP-9E9990/2020\|EPI_ISL_584271\|20 | ···················· | ···················· | ····················· | ···················· | ···················· | ···················· |
| M | 5124 hCoV-19/England/ALDP-9E99CD/2020\|EPI_ISL_584274\|20 | ···················· | ···················· | ····················· | ···················· | ···················· | ···················· |
| M | 5125 hCoV-19/England/ALDP-9E99FA/2020\|EPI_ISL_584277\|20 | ···················· | ···················· | ····················· | ···················· | ···················· | ···················· |
| M | 5126 hCoV-19/England/ALDP-9E9A06/2020\|EPI_ISL_584278\|20 | ···················· | ···················· | ····················· | ···················· | ···················· | ···················· |
| M | 5127 hCoV-19/England/ALDP-9E9A24/2020\|EPI_ISL_584280\|20 | ···················· | ···················· | ····················· | ···················· | ···················· | ···················· |
| M | 5128 hCoV-19/England/ALDP-9E9A33/2020\|EPI_ISL_584281\|20 | ···················· | ···················· | ····················· | ···················· | ···················· | ···················· |
| M | 5129 hCoV-19/England/ALDP-9E9A42/2020\|EPI_ISL_584282\|20 | ···················· | ···················· | ····················· | ···················· | ···················· | ···················· |
| M | 5132 hCoV-19/Scotland/QEUH-A00DF0/2020\|EPI_ISL_585576\|2 | ···················· | ···················· | ····················· | ···················· | ···················· | ···················· |
| M | 5133 hCoV-19/Scotland/QEUH-A00E0C/2020\|EPI_ISL_585577\|2 | ···················· | ···················· | ····················· | ···················· | ·····T·············· | ···················· |
| M | 5134 hCoV-19/Scotland/QEUH-A00E2A/2020\|EPI_ISL_585579\|2 | ···················· | ···················· | ····················· | ···················· | ·····T·············· | ···················· |
| M | 5135 hCoV-19/England/ALDP-A2E14B/2020\|EPI_ISL_586800\|20 | ···················· | ···················· | ····················· | ···················· | ···················· | ···················· |
| M | 5136 hCoV-19/England/ALDP-A2DF42/2020\|EPI_ISL_586801\|20 | ···················· | ···················· | ····················· | ···················· | ···················· | ···················· |
| M | 5137 hCoV-19/England/ALDP-A2E63D/2020\|EPI_ISL_586802\|20 | ···················· | ···················· | ····················· | ···················· | ···················· | ···················· |
| M | 5138 hCoV-19/England/ALDP-A2DC0F/2020\|EPI_ISL_586803\|20 | ···················· | ···················· | ····················· | ···················· | ···················· | ···················· |
| M | 5139 hCoV-19/England/ALDP-A2D0B8/2020\|EPI_ISL_586804\|20 | ···················· | ···················· | ····················· | ···················· | ···················· | ···················· |
| M | 5140 hCoV-19/England/ALDP-A2D8FC/2020\|EPI_ISL_586805\|20 | ···················· | ·A·················· | ····················· | ···················· | ···················· | ··A················· |
| M | 5141 hCoV-19/England/ALDP-A2D70E/2020\|EPI_ISL_586806\|20 | ···················· | ···················· | ····················· | ···················· | ···················· | ···················· |
| M | 5142 hCoV-19/England/ALDP-A2D7B3/2020\|EPI_ISL_586807\|20 | ···················· | ···················· | ····················· | ···················· | ···················· | ···················· |
| M | 5143 hCoV-19/England/ALDP-A2E4BB/2020\|EPI_ISL_586809\|20 | ···················· | ···················· | ····················· | ···················· | ···················· | ···················· |
| M | 5144 hCoV-19/England/ALDP-A2D4E9/2020\|EPI_ISL_586810\|20 | ···················· | ···················· | ····················· | ···················· | ···················· | ···················· |
| M | 5145 hCoV-19/England/ALDP-A2E451/2020\|EPI_ISL_586811\|20 | ···················· | ···················· | ····················· | ···················· | ···················· | ···················· |
| M | 5146 hCoV-19/England/ALDP-A2D1B5/2020\|EPI_ISL_586812\|20 | ···················· | ···················· | ····················· | ···················· | ···················· | ···················· |
| M | 5147 hCoV-19/England/ALDP-A2E688/2020\|EPI_ISL_586813\|20 | ···················· | ···················· | ····················· | ···················· | ···················· | ···················· |
| M | 5148 hCoV-19/England/ALDP-A2D391/2020\|EPI_ISL_586814\|20 | ···················· | ···················· | ····················· | ···················· | ···················· | ···················· |
| M | 5149 hCoV-19/England/ALDP-A2E1F0/2020\|EPI_ISL_586815\|20 | ···················· | ···················· | ····················· | ···················· | ···················· | ···················· |
| M | 5150 hCoV-19/England/ALDP-A2D188/2020\|EPI_ISL_586816\|20 | ···················· | ···················· | ····················· | ···················· | ···················· | ···················· |
| M | 5151 hCoV-19/England/ALDP-A2E187/2020\|EPI_ISL_586817\|20 | ···················· | ···················· | ····················· | ···················· | ···················· | ···················· |
| M | 5152 hCoV-19/England/ALDP-A2D48F/2020\|EPI_ISL_586818\|20 | ···················· | ···················· | ····················· | ···················· | ···················· | ···················· |
| M | 5153 hCoV-19/England/ALDP-A2E828/2020\|EPI_ISL_586819\|20 | ···················· | ···················· | ····················· | ···················· | ···················· | ···················· |
| M | 5154 hCoV-19/England/QEUH-9F8970/2020\|EPI_ISL_589000\|20 | ···················· | ···················· | ····················· | ···················· | ···················· | ···················· |
| M | 5155 hCoV-19/England/QEUH-9F820C/2020\|EPI_ISL_589001\|20 | ···················· | ···················· | ····················· | ···················· | ···················· | ···················· |
| M | 5156 hCoV-19/England/QEUH-9F6365/2020\|EPI_ISL_589002\|20 | ···················· | ···················· | ····················· | ···················· | ···················· | ···················· |
| M | 5157 hCoV-19/England/QEUH-9F8E35/2020\|EPI_ISL_589003\|20 | ···················· | ···················· | ····················· | ···················· | ···T················ | ···················· |
| M | 5158 hCoV-19/England/QEUH-9F6277/2020\|EPI_ISL_589004\|20 | ···················· | ···················· | ····················· | ···················· | ···················· | ···················· |
| M | 5159 hCoV-19/England/QEUH-9F606E/2020\|EPI_ISL_589005\|20 | ···················· | ···················· | ····················· | ···················· | ···················· | ···················· |
| M | 5160 hCoV-19/England/QEUH-9F8011/2020\|EPI_ISL_589006\|20 | ···················· | ···················· | ····················· | ···················· | ···················· | ···················· |
| M | 5161 hCoV-19/England/QEUH-9F62C2/2020\|EPI_ISL_589007\|20 | ···················· | ···················· | ····················· | ···················· | ···················· | ···················· |
| M | 5162 hCoV-19/England/QEUH-9F8BA7/2020\|EPI_ISL_589008\|20 | ···················· | ···················· | ····················· | ···················· | ···················· | ···················· |
| M | 5163 hCoV-19/England/QEUH-9F8819/2020\|EPI_ISL_589009\|20 | ···················· | ···················· | ····················· | ···················· | ···················· | ···················· |
| M | 5164 hCoV-19/England/QEUH-9F56A9/2020\|EPI_ISL_589010\|20 | ···················· | ···················· | ····················· | ···················· | ···················· | ···················· |
| M | 5165 hCoV-19/England/QEUH-9F8CB3/2020\|EPI_ISL_589011\|20 | ···················· | ···················· | ····················· | ···················· | ···················· | ···················· |
| M | 5166 hCoV-19/England/QEUH-9F6462/2020\|EPI_ISL_589013\|20 | ···················· | ···················· | ····················· | ···················· | ···················· | ···················· |
| M | 5167 hCoV-19/England/QEUH-9F8CD1/2020\|EPI_ISL_589014\|20 | ···················· | ···················· | ····················· | ···················· | ···················· | ···················· |
| M | 5168 hCoV-19/England/QEUH-9F906B/2020\|EPI_ISL_589016\|20 | ···················· | ···················· | ····················· | ···················· | ···················· | ···················· |
| M | 5169 hCoV-19/England/MILK-9F26AC/2020\|EPI_ISL_589017\|20 | ···················· | ···················· | ····················· | ···················· | ···················· | ···················· |
| M | 5170 hCoV-19/England/MILK-9F27D6/2020\|EPI_ISL_589018\|20 | ···················· | ···················· | ····················· | ···················· | ···················· | ···················· |
| M | 5171 hCoV-19/England/MILK-9F267F/2020\|EPI_ISL_589030\|20 | ···················· | ···················· | ····················· | ···················· | ···················· | ···················· |
| M | 5172 hCoV-19/England/QEUH-9F965A/2020\|EPI_ISL_589031\|20 | ···················· | ···················· | ····················· | ···················· | ···················· | ···················· |
| M | 5173 hCoV-19/England/QEUH-9F9890/2020\|EPI_ISL_589032\|20 | ···················· | ···················· | ····················· | ···················· | ···················· | ···················· |
| M | 5174 hCoV-19/England/QEUH-9F956C/2020\|EPI_ISL_589061\|20 | ···················· | ···················· | ····················· | ···················· | ···················· | ···················· |
| M | 5175 hCoV-19/England/MILK-9F2590/2020\|EPI_ISL_589063\|20 | ···················· | ···················· | ····················· | ···················· | ···················· | ···················· |
| M | 5176 hCoV-19/England/QEUH-9F9A7C/2020\|EPI_ISL_589064\|20 | ···················· | ·A·················· | ····················· | ···················· | ···················· | ··A················· |
| M | 5177 hCoV-19/England/QEUH-9F7BA8/2020\|EPI_ISL_589065\|20 | ···················· | ···················· | ····················· | ···················· | ···T················ | ···················· |
| M | 5178 hCoV-19/England/QEUH-9F9863/2020\|EPI_ISL_589066\|20 | ···················· | ···················· | ····················· | ···················· | ·····T·············· | ···················· |
| M | 5179 hCoV-19/Wales/MILK-9F2949/2020\|EPI_ISL_589067\|2020 | ···················· | ···················· | ····················· | ···················· | ···················· | ···················· |
| M | 5180 hCoV-19/England/QEUH-9F7443/2020\|EPI_ISL_589068\|20 | ···················· | ···················· | ····················· | ···················· | ···················· | ···················· |
| M | 5181 hCoV-19/England/QEUH-9F7E27/2020\|EPI_ISL_589069\|20 | ···················· | ···················· | ····················· | ···················· | ···················· | ···················· |
| M | 5182 hCoV-19/Wales/MILK-9F2A73/2020\|EPI_ISL_589070\|2020 | ···················· | ···················· | ····················· | ···················· | ···················· | ···················· |
| M | 5183 hCoV-19/England/QEUH-9F7A8D/2020\|EPI_ISL_589072\|20 | ···················· | ···················· | ····················· | ···················· | ···················· | ···················· |
| M | 5184 hCoV-19/England/QEUH-9F9748/2020\|EPI_ISL_589073\|20 | ···················· | ···················· | ····················· | ···················· | ···················· | ···················· |
| M | 5185 hCoV-19/England/MILK-9F2660/2020\|EPI_ISL_589074\|20 | ···················· | ···················· | ····················· | ···················· | ···················· | ···················· |
| M | 5186 hCoV-19/England/MILK-9F26D9/2020\|EPI_ISL_589075\|20 | ···················· | ·A·················· | ····················· | ···················· | ···················· | ···················· |
| M | 5187 hCoV-19/England/QEUH-9F961E/2020\|EPI_ISL_589076\|20 | ···················· | ···················· | ····················· | ···················· | ···················· | ···················· |
| M | 5188 hCoV-19/England/MILK-9F2053/2020\|EPI_ISL_589077\|20 | ···················· | ···················· | ····················· | ···················· | ···················· | ···················· |
| M | 5189 hCoV-19/England/QEUH-9F7B6C/2020\|EPI_ISL_589078\|20 | ···················· | ···················· | ····················· | ···················· | ···T················ | ···················· |
| M | 5190 hCoV-19/England/MILK-9F232D/2020\|EPI_ISL_589080\|20 | ···················· | ···················· | ····················· | ···················· | ···················· | ···················· |
| M | 5191 hCoV-19/England/MILK-9F20EA/2020\|EPI_ISL_589081\|20 | ···················· | ···················· | ····················· | ···················· | ···················· | ···················· |
| M | 5192 hCoV-19/England/QEUH-9F7B4E/2020\|EPI_ISL_589082\|20 | ···················· | ···················· | ····················· | ···················· | ···················· | ···················· |
| M | 5193 hCoV-19/England/QEUH-9F7504/2020\|EPI_ISL_589084\|20 | ···················· | ···················· | ····················· | ···················· | ···················· | ···················· |
| M | 5194 hCoV-19/Wales/MILK-9F23E1/2020\|EPI_ISL_589086\|2020 | ···················· | ···················· | ····················· | ···················· | ···················· | ···················· |
| M | 5195 hCoV-19/England/QEUH-9F7C2D/2020\|EPI_ISL_589087\|20 | ···················· | ···················· | ····················· | ···················· | ···················· | ···················· |
| M | 5196 hCoV-19/England/QEUH-9F964B/2020\|EPI_ISL_589088\|20 | ···················· | ···················· | ····················· | ···················· | ···················· | ···················· |
| M | 5197 hCoV-19/England/QEUH-9F7B02/2020\|EPI_ISL_589089\|20 | ···················· | ···················· | ····················· | ···················· | ···················· | ···················· |
| M | 5198 hCoV-19/England/MILK-9F209F/2020\|EPI_ISL_589090\|20 | ···················· | ···················· | ····················· | ···················· | ···················· | ···················· |
| M | 5199 hCoV-19/England/MILK-9F268E/2020\|EPI_ISL_589091\|20 | ···················· | ···················· | ····················· | ···················· | ···················· | ···················· |
| M | 5200 hCoV-19/England/QEUH-9F748F/2020\|EPI_ISL_589092\|20 | ···················· | ·A·················· | ····················· | ···················· | ···················· | ···················· |
| M | 5201 hCoV-19/England/QEUH-9F9775/2020\|EPI_ISL_589093\|20 | ···················· | ·A·················· | ····················· | ···················· | ···················· | ··A················· |
| M | 5202 hCoV-19/England/QEUH-9F9845/2020\|EPI_ISL_589094\|20 | ···················· | ···················· | ····················· | ···················· | ···················· | ···················· |
| M | 5203 hCoV-19/England/MILK-9F25CD/2020\|EPI_ISL_589095\|20 | ···················· | ···················· | ····················· | ···················· | ···················· | ···················· |
| M | 5204 hCoV-19/England/QEUH-9F7BD5/2020\|EPI_ISL_589096\|20 | ···················· | ···················· | ····················· | ···················· | ···················· | ···················· |

| | | | | | | | |
|---|---|---|---|---|---|---|---|
| M | 5205 hCoV-19/England/MILK-9F235A/2020\|EPI_ISL_589097\|20 | ···················· | ···················· | ···················· | ···················· | ···················· | ···················· |
| M | 5206 hCoV-19/England/MILK-9F217E/2020\|EPI_ISL_589098\|20 | ···················· | ···················· | ···················· | ···················· | ···················· | ···················· |
| M | 5207 hCoV-19/England/MILK-9F2114/2020\|EPI_ISL_589099\|20 | ···················· | ···················· | ···················· | ···················· | ···················· | ···················· |
| M | 5208 hCoV-19/England/QEUH-9CEBC8/2020\|EPI_ISL_590017\|20 | ···················· | ·A·················· | ···················· | ···················· | ···················· | ···················· |
| M | 5209 hCoV-19/England/QEUH-9CF58D/2020\|EPI_ISL_590018\|20 | ···················· | ···················· | ···················· | ···················· | ···················· | ···················· |
| M | 5210 hCoV-19/England/QEUH-9CEC89/2020\|EPI_ISL_590043\|20 | ···················· | ···················· | ···················· | ···················· | ···················· | ···················· |
| M | 5211 hCoV-19/England/QEUH-9CEB31/2020\|EPI_ISL_590044\|20 | ···················· | ···················· | ···················· | ···················· | ···················· | ···················· |
| M | 5212 hCoV-19/England/QEUH-9CEC01/2020\|EPI_ISL_590045\|20 | ···················· | ···················· | ···················· | ···················· | ···················· | ···················· |
| M | 5213 hCoV-19/England/QEUH-9CF3ED/2020\|EPI_ISL_590048\|20 | ···················· | ···················· | ···················· | ···················· | ···················· | ···················· |
| M | 5214 hCoV-19/England/QEUH-9CEF80/2020\|EPI_ISL_590049\|20 | ···················· | ···················· | ···················· | ···················· | ···················· | ···················· |
| M | 5215 hCoV-19/England/QEUH-9CE20F/2020\|EPI_ISL_590050\|20 | ···················· | ···················· | ···················· | ···················· | ···················· | ···················· |
| M | 5216 hCoV-19/England/QEUH-9CE4DC/2020\|EPI_ISL_590051\|20 | ···················· | ·A·················· | ···················· | ···················· | ···················· | ···················· |
| M | 5217 hCoV-19/England/QEUH-9CF16B/2020\|EPI_ISL_590052\|20 | ···················· | ···················· | ···················· | ···················· | ···················· | ···················· |
| M | 5218 hCoV-19/England/QEUH-9CEDC2/2020\|EPI_ISL_590053\|20 | ···················· | ···················· | ···················· | ···················· | ···················· | ···················· |
| M | 5219 hCoV-19/England/QEUH-9CF5BA/2020\|EPI_ISL_590054\|20 | ···················· | ·A·················· | ···················· | ···················· | ···················· | ···················· |
| M | 5220 hCoV-19/England/QEUH-9CF59C/2020\|EPI_ISL_590055\|20 | ···················· | ···················· | ···················· | ···················· | ···················· | ···················· |
| M | 5221 hCoV-19/England/QEUH-9CEE83/2020\|EPI_ISL_590056\|20 | ···················· | ···················· | ···················· | ···················· | ···················· | ···················· |
| M | 5222 hCoV-19/England/QEUH-9CE57F/2020\|EPI_ISL_590057\|20 | ···················· | ···················· | ···················· | ···················· | ···················· | ···················· |
| M | 5223 hCoV-19/England/QEUH-9CF5C9/2020\|EPI_ISL_590058\|20 | ···················· | ···················· | ···················· | ···················· | ···················· | ···················· |
| M | 5224 hCoV-19/England/QEUH-9CEF71/2020\|EPI_ISL_590059\|20 | ···················· | ···················· | ···················· | ···················· | ···················· | ···················· |
| M | 5225 hCoV-19/England/QEUH-9CF963/2020\|EPI_ISL_590060\|20 | ···················· | ···················· | ···················· | ···················· | ···················· | ···················· |
| M | 5226 hCoV-19/England/QEUH-9CE6F4/2020\|EPI_ISL_590061\|20 | ···················· | ·A·················· | ···················· | ···················· | ···················· | ···················· |
| M | 5227 hCoV-19/England/QEUH-9CEFDB/2020\|EPI_ISL_590062\|20 | ···················· | ···················· | ···················· | ···················· | ···················· | ···················· |
| M | 5228 hCoV-19/England/QEUH-9CE454/2020\|EPI_ISL_590064\|20 | ···················· | ···················· | ···················· | ···················· | ···················· | ···················· |
| M | 5229 hCoV-19/Wales/QEUH-9CF2B3/2020\|EPI_ISL_590065\|2020 | ···················· | ···················· | ···················· | ···················· | ···················· | ···················· |
| M | 5230 hCoV-19/England/QEUH-9CE463/2020\|EPI_ISL_590066\|20 | ···················· | ·A·················· | ···················· | ···················· | ···················· | ···················· |
| M | 5231 hCoV-19/England/QEUH-9CF6B7/2020\|EPI_ISL_590067\|20 | ···················· | ···················· | ···················· | ···················· | ···················· | ···················· |
| M | 5232 hCoV-19/England/QEUH-9CF004/2020\|EPI_ISL_590069\|20 | ···················· | ······C············· | ···················· | ···················· | ···················· | ···················· |
| M | 5233 hCoV-19/England/QEUH-9CED59/2020\|EPI_ISL_590070\|20 | ···················· | ···················· | ···················· | ···················· | ···················· | ···················· |
| M | 5234 hCoV-19/England/QEUH-9CED4A/2020\|EPI_ISL_590071\|20 | ···················· | ···················· | ···················· | ···················· | ···················· | ···················· |
| M | 5235 hCoV-19/England/QEUH-9CE1C6/2020\|EPI_ISL_590072\|20 | ···················· | ···················· | ···················· | ···················· | ···················· | ···················· |
| M | 5236 hCoV-19/England/QEUH-9CF57E/2020\|EPI_ISL_590073\|20 | ···················· | ···················· | ···················· | ···················· | ···················· | ···················· |
| M | 5237 hCoV-19/England/QEUH-9CF3CF/2020\|EPI_ISL_590074\|20 | ···················· | ···················· | ···················· | ···················· | ···················· | ···················· |
| M | 5238 hCoV-19/England/QEUH-9CE779/2020\|EPI_ISL_590076\|20 | ···················· | ···················· | ···················· | ···················· | ···················· | ···················· |
| M | 5239 hCoV-19/England/QEUH-9CF17A/2020\|EPI_ISL_590077\|20 | ···················· | ···················· | ···················· | ···················· | ···················· | ···················· |
| M | 5240 hCoV-19/England/QEUH-9CE73D/2020\|EPI_ISL_590078\|20 | ···················· | ···················· | ···················· | ···················· | ···················· | ···················· |
| M | 5241 hCoV-19/England/QEUH-9CE80D/2020\|EPI_ISL_590080\|20 | ···················· | ···················· | ···················· | ···················· | ···················· | ···················· |
| M | 5242 hCoV-19/England/QEUH-9CE71F/2020\|EPI_ISL_590081\|20 | ···················· | ···················· | ···················· | ···················· | ···················· | ···················· |
| M | 5243 hCoV-19/England/QEUH-9CF65D/2020\|EPI_ISL_590082\|20 | ···················· | ···················· | ···················· | ···················· | ···················· | ···················· |
| M | 5244 hCoV-19/England/QEUH-9CF08C/2020\|EPI_ISL_590083\|20 | ···················· | ···················· | ···················· | ···················· | ···················· | ···················· |
| M | 5245 hCoV-19/England/QEUH-9CEC7A/2020\|EPI_ISL_590084\|20 | ···················· | ···················· | ···················· | ···················· | ······T············· | ···················· |
| M | 5246 hCoV-19/England/QEUH-9CE74C/2020\|EPI_ISL_590085\|20 | ···················· | ···················· | ···················· | ···················· | ···················· | ···················· |
| M | 5247 hCoV-19/USA/UT-UPHL-04309_2/2020\|EPI_ISL_591300\|20 | ···················· | ···················· | ···················· | ···················· | ···················· | ···················· |
| M | 5248 hCoV-19/USA/UT-UPHL-04311_2/2020\|EPI_ISL_591302\|20 | ···················· | ···················· | ···················· | ···················· | ···················· | ···················· |
| M | 5249 hCoV-19/USA/UT-UPHL-04312_2/2020\|EPI_ISL_591303\|20 | ···················· | ···················· | ···················· | ···················· | ···················· | ···················· |
| M | 5250 hCoV-19/USA/UT-UPHL-04313_2/2020\|EPI_ISL_591304\|20 | ···················· | ···················· | ···················· | ···················· | ···················· | ···················· |
| M | 5251 hCoV-19/USA/UT-UPHL-04314_2/2020\|EPI_ISL_591305\|20 | ···················· | ···················· | ···················· | ···················· | ···················· | ···················· |
| M | 5252 hCoV-19/USA/UT-UPHL-04318_2/2020\|EPI_ISL_591308\|20 | ···················· | ·A·················· | ···················· | ···················· | ···················· | ···················· |
| M | 5253 hCoV-19/USA/UT-UPHL-04319_2/2020\|EPI_ISL_591309\|20 | ···················· | ···················· | ···················· | ···················· | ···················· | ···················· |
| M | 5254 hCoV-19/USA/UT-UPHL-04341_2/2020\|EPI_ISL_591315\|20 | ···················· | ···················· | ···················· | ···················· | ···················· | ···················· |
| M | 5255 hCoV-19/Italy/TUS-UOS-2/2020\|EPI_ISL_591326\|2020-0 | ···················· | ···················· | ···················· | ···················· | ···················· | ···················· |
| M | 5256 hCoV-19/Italy/TUS-UOS-10/2020\|EPI_ISL_591327\|2020- | ···················· | ···················· | ···················· | ···················· | ···················· | ···················· |
| M | 5257 hCoV-19/Italy/TUS-UOS-11/2020\|EPI_ISL_591328\|2020- | ···················· | ···················· | ···················· | ···················· | ···················· | ···················· |
| M | 5258 hCoV-19/Italy/TUS-UOS-18/2020\|EPI_ISL_591329\|2020- | ···················· | ···················· | ···················· | ···················· | ···················· | ···················· |
| M | 5259 hCoV-19/Japan/IC-0002/2020\|EPI_ISL_591348\|2020-03 | ···················· | ·A·················· | ···················· | ···················· | ···················· | ···················· |
| M | 5260 hCoV-19/Japan/IC-0003/2020\|EPI_ISL_591349\|2020-03 | ···················· | ···················· | ···················· | ···················· | ···················· | ···················· |
| M | 5261 hCoV-19/Japan/IC-0029/2020\|EPI_ISL_591373\|2020-05 | ···················· | ···················· | ···················· | ···················· | ···················· | ···················· |
| M | 5262 hCoV-19/Japan/IC-0030/2020\|EPI_ISL_591374\|2020-05 | ···················· | ···················· | ···················· | ···················· | ···················· | ···················· |
| M | 5263 hCoV-19/Japan/IC-0033/2020\|EPI_ISL_591375\|2020-06 | ···················· | ···················· | ···················· | ···················· | ···················· | ···················· |
| M | 5264 hCoV-19/Japan/IC-0034/2020\|EPI_ISL_591376\|2020-03 | ···················· | ···················· | ···················· | ···················· | ···················· | ···················· |
| M | 5265 hCoV-19/Japan/IC-0035/2020\|EPI_ISL_591377\|2020-04 | ···················· | ···················· | ···················· | ···················· | ···················· | ···················· |
| M | 5266 hCoV-19/Japan/IC-0036/2020\|EPI_ISL_591378\|2020-04 | ···················· | ···················· | ···················· | ···················· | ···················· | ···················· |
| M | 5267 hCoV-19/Japan/IC-0037/2020\|EPI_ISL_591379\|2020-05 | ···················· | ···················· | ···················· | ···················· | ···················· | ···················· |
| M | 5268 hCoV-19/Japan/IC-0038/2020\|EPI_ISL_591380\|2020-05 | ···················· | ···················· | ···················· | ···················· | ···················· | ···················· |
| M | 5269 hCoV-19/Japan/IC-0039/2020\|EPI_ISL_591381\|2020-05 | ···················· | ···················· | ···················· | ···················· | ···················· | ···················· |
| M | 5270 hCoV-19/Japan/IC-0040/2020\|EPI_ISL_591382\|2020-05 | ···················· | ···················· | ···················· | ···················· | ···················· | ···················· |
| M | 5271 hCoV-19/Japan/IC-0041/2020\|EPI_ISL_591383\|2020-05 | ···················· | ···················· | ···················· | ···················· | ···················· | ···················· |
| M | 5272 hCoV-19/Japan/IC-0042/2020\|EPI_ISL_591384\|2020-05 | ···················· | ···················· | ···················· | ···················· | ···················· | ···················· |
| M | 5273 hCoV-19/Japan/IC-0043/2020\|EPI_ISL_591385\|2020-05 | ···················· | ···················· | ···················· | ···················· | ···················· | ···················· |
| M | 5274 hCoV-19/Japan/IC-0044/2020\|EPI_ISL_591386\|2020-05 | ···················· | ···················· | ···················· | ···················· | ···················· | ···················· |
| M | 5275 hCoV-19/Japan/IC-0045/2020\|EPI_ISL_591387\|2020-05 | ···················· | ···················· | ···················· | ···················· | ···················· | ···················· |
| M | 5276 hCoV-19/Japan/IC-0046/2020\|EPI_ISL_591388\|2020-05 | ···················· | ···················· | ···················· | ···················· | ···················· | ···················· |
| M | 5277 hCoV-19/Japan/IC-0047/2020\|EPI_ISL_591389\|2020-05 | ···················· | ···················· | ···················· | ···················· | ···················· | ···················· |
| M | 5278 hCoV-19/Japan/IC-0048/2020\|EPI_ISL_591390\|2020-05 | ···················· | ···················· | ···················· | ···················· | ···················· | ···················· |
| M | 5279 hCoV-19/Japan/IC-0049/2020\|EPI_ISL_591391\|2020-06 | ···················· | ···················· | ···················· | ···················· | ···················· | ···················· |
| M | 5280 hCoV-19/Japan/IC-0050/2020\|EPI_ISL_591392\|2020-06 | ···················· | ···················· | ···················· | ···················· | ···················· | ···················· |
| M | 5281 hCoV-19/Japan/IC-0051/2020\|EPI_ISL_591393\|2020-06 | ···················· | ···················· | ···················· | ···················· | ···················· | ···················· |
| M | 5282 hCoV-19/Japan/IC-0052/2020\|EPI_ISL_591394\|2020-06 | ···················· | ···················· | ···················· | ···················· | ···················· | ···················· |
| M | 5283 hCoV-19/Japan/IC-0053/2020\|EPI_ISL_591395\|2020-06 | ···················· | ···················· | ···················· | ···················· | ···················· | ···················· |
| M | 5284 hCoV-19/Japan/IC-0054/2020\|EPI_ISL_591396\|2020-06 | ···················· | ···················· | ···················· | ···················· | ···················· | ···················· |

| | | | | | | | |
|---|---|---|---|---|---|---|---|
| M | 5285 hCoV-19/Japan/IC-0055/2020\|EPI_ISL_591397\|2020-06 | .................... | .................... | .................... | .................... | .................... | .................... |
| M | 5286 hCoV-19/Japan/IC-0056/2020\|EPI_ISL_591398\|2020-06 | .................... | .................... | .................... | .................... | .................... | .................... |
| M | 5287 hCoV-19/Japan/IC-0057/2020\|EPI_ISL_591399\|2020-06 | .................... | .................... | .................... | .................... | .................... | .................... |
| M | 5288 hCoV-19/France/IDF-9144/2020\|EPI_ISL_593910\|2020-0 | .................... | .................... | .................... | .................... | .................... | .................... |
| M | 5289 hCoV-19/France/IDF-9146/2020\|EPI_ISL_593911\|2020-0 | .................... | .................... | .................... | .................... | .................... | .................... |
| M | 5290 hCoV-19/France/IDF-9147/2020\|EPI_ISL_593912\|2020-0 | .................... | .................... | .................... | .................... | .................... | .................... |
| M | 5291 hCoV-19/France/IDF-9155/2020\|EPI_ISL_593913\|2020-0 | .................... | .................... | .................... | .................... | .................... | .................... |
| M | 5292 hCoV-19/France/IDF-9157/2020\|EPI_ISL_593914\|2020-0 | .................... | .................... | .................... | .................... | .................... | .................... |
| M | 5293 hCoV-19/France/IDF-9163/2020\|EPI_ISL_593915\|2020-0 | .................... | .................... | .................... | .................... | .................... | .................... |
| M | 5294 hCoV-19/France/IDF-9164/2020\|EPI_ISL_593916\|2020-0 | .................... | .................... | .................... | .................... | .................... | .................... |
| M | 5295 hCoV-19/France/IDF-9171/2020\|EPI_ISL_593917\|2020-0 | .................... | .A.................. | .................... | .................... | .................... | .................... |
| M | 5296 hCoV-19/France/IDF-9172/2020\|EPI_ISL_593918\|2020-0 | .................... | .................... | .................... | .................... | .................... | .................... |
| M | 5297 hCoV-19/France/IDF-9175/2020\|EPI_ISL_593919\|2020-0 | .................... | .................... | .................... | .................... | .................... | .................... |
| M | 5298 hCoV-19/France/IDF-9177/2020\|EPI_ISL_593920\|2020-0 | .................... | .................... | .................... | .................... | .................... | .................... |
| M | 5299 hCoV-19/France/IDF-9183/2020\|EPI_ISL_593921\|2020-0 | .................... | .................... | .................... | .................... | .................... | .................... |
| M | 5300 hCoV-19/France/IDF-9190/2020\|EPI_ISL_593922\|2020-0 | .................... | .................... | .................... | .................... | .................... | .................... |
| M | 5301 hCoV-19/France/IDF-9191/2020\|EPI_ISL_593923\|2020-0 | .................... | .................... | .................... | .................... | .................... | .................... |
| M | 5302 hCoV-19/France/IDF-9128/2020\|EPI_ISL_593924\|2020-0 | .................... | .................... | .................... | .................... | .................... | .................... |
| M | 5303 hCoV-19/France/IDF-9131/2020\|EPI_ISL_593925\|2020-0 | .................... | .................... | .................... | .................... | .................... | .................... |
| M | 5304 hCoV-19/France/IDF-9136/2020\|EPI_ISL_593926\|2020-0 | .................... | .................... | .................... | .................... | .................... | .................... |
| M | 5305 hCoV-19/France/IDF-9138/2020\|EPI_ISL_593927\|2020-0 | .................... | .................... | .................... | .................... | .................... | .................... |
| M | 5306 hCoV-19/France/IDF-9140/2020\|EPI_ISL_593928\|2020-0 | .................... | .................... | .................... | .................... | .................... | .................... |
| M | 5307 hCoV-19/France/IDF-9149/2020\|EPI_ISL_593929\|2020-0 | .................... | .................... | .................... | .................... | .................... | .................... |
| M | 5308 hCoV-19/France/IDF-9199/2020\|EPI_ISL_593932\|2020-0 | .................... | .................... | .................... | .................... | .................... | .................... |
| M | 5309 hCoV-19/France/IDF-9202/2020\|EPI_ISL_593933\|2020-0 | .................... | .................... | .................... | .................... | .................... | .................... |
| M | 5310 hCoV-19/France/IDF-9230/2020\|EPI_ISL_593934\|2020-0 | .................... | .................... | .................... | .................... | .................... | .................... |
| M | 5311 hCoV-19/France/IDF-9320/2020\|EPI_ISL_593935\|2020-0 | .................... | .................... | .................... | .................... | .................... | .................... |
| M | 5312 hCoV-19/France/CVL-9321/2020\|EPI_ISL_593936\|2020-1 | .................... | .................... | .................... | .................... | .................... | .................... |
| M | 5313 hCoV-19/USA/DE-DHSS-F918037/2020\|EPI_ISL_593938\|20 | .................... | .................... | .................... | .................... | .................... | .................... |
| M | 5314 hCoV-19/USA/DE-DHSS-F918049/2020\|EPI_ISL_593940\|20 | .................... | .................... | .................... | .................... | .................... | .................... |
| M | 5315 hCoV-19/USA/DE-DHSS-F918170/2020\|EPI_ISL_593945\|20 | .................... | .................... | .................... | .................... | .................... | .................... |
| M | 5316 hCoV-19/USA/DE-DHSS-F918349/2020\|EPI_ISL_593949\|20 | .................... | .................... | .................... | .................... | .................... | .................... |
| M | 5317 hCoV-19/USA/DE-DHSS-F918376/2020\|EPI_ISL_593950\|20 | .................... | .................... | .................... | .................... | .................... | .................... |
| M | 5318 hCoV-19/USA/DE-DHSS-F918378/2020\|EPI_ISL_593951\|20 | .................... | .................... | .................... | .................... | .................... | .................... |
| M | 5319 hCoV-19/USA/DE-DHSS-F918384/2020\|EPI_ISL_593952\|20 | .................... | .................... | .................... | .................... | .................... | .................... |
| M | 5320 hCoV-19/USA/DE-DHSS-F918388/2020\|EPI_ISL_593953\|20 | .................... | .................... | .................... | .................... | .................... | .................... |
| M | 5321 hCoV-19/USA/DE-DHSS-F918400/2020\|EPI_ISL_593955\|20 | .................... | .................... | .................... | .................... | .................... | .................... |
| M | 5322 hCoV-19/USA/DE-DHSS-F918402/2020\|EPI_ISL_593956\|20 | .................... | .................... | .................... | .................... | .................... | .................... |
| M | 5323 hCoV-19/USA/DE-DHSS-F918416/2020\|EPI_ISL_593957\|20 | .................... | .................... | .................... | .................... | .................... | .................... |
| M | 5324 hCoV-19/USA/DE-DHSS-F918418/2020\|EPI_ISL_593958\|20 | .................... | .................... | .................... | .................... | .................... | .................... |
| M | 5325 hCoV-19/USA/DE-DHSS-F918443/2020\|EPI_ISL_593963\|20 | .................... | .................... | .................... | .................... | .................... | .................... |
| M | 5326 hCoV-19/USA/DE-DHSS-F918445/2020\|EPI_ISL_593965\|20 | .................... | .................... | .................... | .................... | .................... | .................... |
| M | 5327 hCoV-19/USA/DE-DHSS-F918451/2020\|EPI_ISL_593966\|20 | .................... | .................... | .................... | .................... | .................... | .................... |
| M | 5328 hCoV-19/USA/DE-DHSS-F918455/2020\|EPI_ISL_593967\|20 | .................... | .................... | .................... | .................... | .................... | .................... |
| M | 5329 hCoV-19/USA/DE-DHSS-F918464/2020\|EPI_ISL_593969\|20 | .................... | .................... | .................... | .................... | .................... | .................... |
| M | 5330 hCoV-19/USA/DE-DHSS-F918466/2020\|EPI_ISL_593970\|20 | .................... | .................... | .................... | .................... | .................... | .................... |
| M | 5331 hCoV-19/USA/DE-DHSS-F918476/2020\|EPI_ISL_593971\|20 | .................... | .................... | .................... | .................... | .................... | .................... |
| M | 5332 hCoV-19/USA/DE-DHSS-F918493/2020\|EPI_ISL_593973\|20 | .................... | .................... | .................... | .................... | .................... | .................... |
| M | 5333 hCoV-19/USA/DE-DHSS-F918497/2020\|EPI_ISL_593974\|20 | .................... | .................... | .................... | .................... | .................... | .................... |
| M | 5334 hCoV-19/USA/DE-DHSS-F952774/2020\|EPI_ISL_593976\|20 | .................... | .................... | .................... | .................... | .................... | .................... |
| M | 5335 hCoV-19/USA/DE-DHSS-F952974/2020\|EPI_ISL_593979\|20 | .................... | .................... | .................... | .................... | .................... | .................... |
| M | 5336 hCoV-19/USA/DE-DHSS-F994880/2020\|EPI_ISL_593980\|20 | .................... | .A.................. | .................... | .................... | .................... | .................... |
| M | 5337 hCoV-19/USA/DE-DHSS-F996119/2020\|EPI_ISL_593981\|20 | .................... | .A.................. | .................... | .................... | .................... | .................... |
| M | 5338 hCoV-19/England/204221423/2020\|EPI_ISL_593984\|2020 | .................... | .................... | .................... | .................... | .................... | .................... |
| M | 5339 hCoV-19/England/204221434/2020\|EPI_ISL_593985\|2020 | .................... | .................... | .................... | .................... | .................... | .................... |
| M | 5340 hCoV-19/England/204221486/2020\|EPI_ISL_593986\|2020 | .................... | .................... | .................... | .................... | .................... | .................... |
| M | 5341 hCoV-19/England/204221487/2020\|EPI_ISL_593987\|2020 | .................... | .................... | .................... | .................... | .................... | .................... |
| M | 5342 hCoV-19/England/204221490/2020\|EPI_ISL_593989\|2020 | .................... | .................... | .................... | .................... | .................... | .................... |
| M | 5343 hCoV-19/England/204221491/2020\|EPI_ISL_593990\|2020 | .................... | .................... | .................... | .................... | .................... | .................... |
| M | 5344 hCoV-19/England/SHEF-C1FC2/2020\|EPI_ISL_596191\|202 | .................... | .................... | .................... | .................... | .................... | .................... |
| M | 5345 hCoV-19/England/SHEF-C2495/2020\|EPI_ISL_596192\|202 | .................... | .................... | .................... | .................... | .................... | .................... |
| M | 5346 hCoV-19/England/SHEF-C2705/2020\|EPI_ISL_596193\|202 | .................... | .A.................. | .................... | .................... | .................... | .................... |
| M | 5347 hCoV-19/England/SHEF-C42D5/2020\|EPI_ISL_596195\|202 | .................... | .................... | .................... | .................... | .................... | .................... |
| M | 5348 hCoV-19/England/SHEF-C477C/2020\|EPI_ISL_596197\|202 | .................... | .................... | .................... | .................... | .................... | .................... |
| M | 5349 hCoV-19/England/SHEF-C5BD9/2020\|EPI_ISL_596198\|202 | .................... | .................... | .................... | .................... | .................... | .................... |
| M | 5350 hCoV-19/Russia/SPE-RII-33958V/2020\|EPI_ISL_596232\| | .................... | .................... | .................... | .................... | .................... | .................... |
| M | 5351 hCoV-19/Russia/LEN-RII-34268M/2020\|EPI_ISL_596233\| | .................... | .................... | .................... | .................... | .................... | .................... |
| M | 5352 hCoV-19/Russia/LEN-RII-34271M/2020\|EPI_ISL_596234\| | .................... | .................... | .................... | .................... | .................... | .................... |
| M | 5353 hCoV-19/Russia/SPE-RII-35217S/2020\|EPI_ISL_596246\| | .................... | .................... | .................... | .................... | .................... | .................... |
| M | 5354 hCoV-19/Russia/SPE-RII-35397S/2020\|EPI_ISL_596247\| | .................... | .................... | .................... | .................... | .................... | .................... |
| M | 5355 hCoV-19/Russia/SPE-RII-35435S/2020\|EPI_ISL_596248\| | .................... | .................... | .................... | .................... | .................... | .................... |
| M | 5356 hCoV-19/Russia/SPE-RII-35589S/2020\|EPI_ISL_596249\| | .................... | .................... | .................... | .................... | .................... | .................... |
| M | 5357 hCoV-19/Russia/SPE-RII-MH1007S/2020\|EPI_ISL_596250 | .................... | .................... | .................... | .................... | .................... | .................... |
| M | 5358 hCoV-19/Russia/SPE-RII-MH1008S/2020\|EPI_ISL_596251 | .................... | .................... | .................... | .................... | .................... | .................... |
| M | 5359 hCoV-19/Russia/SPE-RII-MH1012S/2020\|EPI_ISL_596252 | .................... | .................... | .................... | .................... | .................... | .................... |
| M | 5360 hCoV-19/Russia/SVE-RII-MH1028S/2020\|EPI_ISL_596253 | .................... | .................... | .................... | .................... | .................... | .................... |
| M | 5361 hCoV-19/Russia/SPE-RII-MH1042S/2020\|EPI_ISL_596254 | .................... | .................... | .................... | .................... | .................... | .................... |
| M | 5362 hCoV-19/Russia/SPE-RII-MH1043S/2020\|EPI_ISL_596255 | .................... | .................... | .................... | .................... | .................... | .................... |
| M | 5363 hCoV-19/Russia/SPE-RII-MH1049S/2020\|EPI_ISL_596256 | .................... | .................... | .................... | .................... | .................... | .................... |
| M | 5364 hCoV-19/Russia/SPE-RII-MH1050S/2020\|EPI_ISL_596257 | .................... | .................... | .................... | .................... | .................... | .................... |

| | | | | | | | | |
|---|---|---|---|---|---|---|---|---|
| M | 5365 | hCoV-19/Russia/SPE-RII-MH1055S/2020\|EPI_ISL_596258 | .................... | .................... | .................... | .................... | .................... | .................... |
| M | 5366 | hCoV-19/Russia/SPE-RII-MH1058S/2020\|EPI_ISL_596259 | .................... | .................... | .................... | .................... | .................... | .................... |
| M | 5367 | hCoV-19/Russia/SPE-RII-MH1059S/2020\|EPI_ISL_596260 | .................... | .................... | .................... | .................... | .................... | .................... |
| M | 5368 | hCoV-19/Russia/SPE-RII-MH1062S/2020\|EPI_ISL_596262 | .................... | .................... | .................... | .................... | .................... | .................... |
| M | 5369 | hCoV-19/Russia/SPE-RII-MH1064S/2020\|EPI_ISL_596263 | .................... | .................... | .................... | .................... | .................... | .................... |
| M | 5370 | hCoV-19/Russia/NVS-RII-MH1087V/2020\|EPI_ISL_596264 | .................... | .................... | .................... | .................... | .................... | .................... |
| M | 5371 | hCoV-19/Russia/NVS-RII-MH1097V/2020\|EPI_ISL_596265 | .................... | .................... | .............A...... | .................... | .................... | .................... |
| M | 5372 | hCoV-19/Russia/NVS-RII-MH1137S/2020\|EPI_ISL_596266 | .................... | .................... | .................... | .................... | .................... | .................... |
| M | 5373 | hCoV-19/Russia/NVS-RII-MH1142S/2020\|EPI_ISL_596267 | .................... | .................... | .................... | .................... | .................... | .................... |
| M | 5374 | hCoV-19/USA/AK-PHL620/2020\|EPI_ISL_602559\|2020-06- | .................... | .................... | .................... | .................... | .................... | .................... |
| M | 5375 | hCoV-19/USA/AK-PHL634/2020\|EPI_ISL_602560\|2020-06- | .................... | .................... | .................... | .................... | .................... | .................... |
| M | 5376 | hCoV-19/USA/AK-PHL783/2020\|EPI_ISL_602561\|2020-07- | .................... | .................... | .................... | .................... | .................... | .................... |
| M | 5377 | hCoV-19/England/ALDP-9E9550/2020\|EPI_ISL_584217\|20 | .................... | .................... | .................... | .................... | .................... | .................... |
| M | 5378 | hCoV-19/England/ALDP-9E9602/2020\|EPI_ISL_584226\|20 | .................... | .................... | .................... | .................... | .................... | .................... |
| M | 5379 | hCoV-19/England/ALDP-9E965D/2020\|EPI_ISL_584230\|20 | .................... | .................... | .................... | .................... | ......T............. | .................... |
| M | 5380 | hCoV-19/England/ALDP-9E966C/2020\|EPI_ISL_584231\|20 | .................... | .A.................. | .................... | .................... | .................... | ...A................ |
| M | 5381 | hCoV-19/England/ALDP-9E967B/2020\|EPI_ISL_584232\|20 | .................... | .................... | .................... | .................... | .................... | .................... |
| M | 5382 | hCoV-19/England/ALDP-9E9699/2020\|EPI_ISL_584233\|20 | .................... | .................... | .................... | .................... | .................... | .................... |
| M | 5383 | hCoV-19/England/ALDP-9E96B7/2020\|EPI_ISL_584234\|20 | .................... | .................... | .................... | .................... | .................... | .................... |
| M | 5384 | hCoV-19/England/ALDP-9E96F3/2020\|EPI_ISL_584238\|20 | .................... | .................... | .................... | .................... | .................... | .................... |
| M | 5385 | hCoV-19/England/ALDP-9E970F/2020\|EPI_ISL_584239\|20 | .................... | .................... | .................... | .................... | .................... | .................... |
| M | 5386 | hCoV-19/England/ALDP-9E973C/2020\|EPI_ISL_584242\|20 | .................... | .................... | .................... | .................... | .................... | .................... |
| M | 5387 | hCoV-19/England/ALDP-9E974B/2020\|EPI_ISL_584243\|20 | .................... | .................... | .................... | .................... | .................... | .................... |
| M | 5388 | hCoV-19/England/ALDP-9E975A/2020\|EPI_ISL_584244\|20 | .................... | .................... | .................... | .................... | .................... | .................... |
| M | 5389 | hCoV-19/England/ALDP-9E9796/2020\|EPI_ISL_584247\|20 | .................... | .................... | .................... | .................... | .................... | .................... |
| M | 5390 | hCoV-19/England/ALDP-9E97C3/2020\|EPI_ISL_584249\|20 | .................... | .................... | .................... | .................... | ....T............... | .................... |
| M | 5391 | hCoV-19/England/ALDP-9E97F0/2020\|EPI_ISL_584252\|20 | .................... | .................... | .................... | .................... | .................... | .................... |
| M | 5392 | hCoV-19/England/ALDP-9E9857/2020\|EPI_ISL_584255\|20 | .................... | .A.................. | .................... | .................... | .................... | ...A................ |
| M | 5393 | hCoV-19/England/ALDP-9E9866/2020\|EPI_ISL_584256\|20 | .................... | .................... | .................... | .................... | .................... | .................... |
| M | 5394 | hCoV-19/England/ALDP-9E9884/2020\|EPI_ISL_584257\|20 | .................... | .................... | .................... | .................... | .................... | .................... |
| M | 5395 | hCoV-19/England/ALDP-9E9893/2020\|EPI_ISL_584258\|20 | .................... | .................... | .................... | .................... | .................... | .................... |
| M | 5396 | hCoV-19/England/ALDP-9E98B1/2020\|EPI_ISL_584260\|20 | .................... | .................... | .................... | .................... | .................... | .................... |
| M | 5397 | hCoV-19/England/ALDP-9E98C0/2020\|EPI_ISL_584261\|20 | .................... | .................... | .................... | .................... | .................... | .................... |
| M | 5398 | hCoV-19/England/ALDP-9E98EE/2020\|EPI_ISL_584262\|20 | .................... | .................... | .................... | .................... | .................... | .................... |
| M | 5399 | hCoV-19/England/ALDP-9E98FD/2020\|EPI_ISL_584263\|20 | .................... | .................... | .................... | .................... | .................... | .................... |
| M | 5400 | hCoV-19/England/ALDP-9E9909/2020\|EPI_ISL_584264\|20 | .................... | .................... | .................... | .................... | .................... | .................... |
| M | 5402 | hCoV-19/England/CAMB-1B667F/2020\|EPI_ISL_584294\|20 | .................... | .................... | .................... | .................... | .................... | .................... |
| M | 5405 | hCoV-19/Scotland/QEUH-9DE907/2020\|EPI_ISL_585502\|2 | .................... | .................... | .................... | .................... | .................... | .................... |
| M | 5406 | hCoV-19/Scotland/QEUH-9DEC0E/2020\|EPI_ISL_585503\|2 | .................... | .................... | .................... | .................... | .................... | .................... |
| M | 5407 | hCoV-19/Scotland/QEUH-9E34B3/2020\|EPI_ISL_585504\|2 | .................... | .................... | .................... | .................... | .................... | .................... |
| M | 5408 | hCoV-19/Scotland/QEUH-9E3538/2020\|EPI_ISL_585506\|2 | .................... | .................... | .................... | .................... | .................... | .................... |
| M | 5409 | hCoV-19/Scotland/QEUH-9E3547/2020\|EPI_ISL_585507\|2 | .................... | .................... | .................... | .................... | .................... | .................... |
| M | 5410 | hCoV-19/Scotland/QEUH-9E3556/2020\|EPI_ISL_585508\|2 | .................... | .................... | .................... | .................... | .................... | .................... |
| M | 5411 | hCoV-19/Scotland/QEUH-9E35DE/2020\|EPI_ISL_585509\|2 | .................... | .................... | .................... | .................... | .................... | .................... |
| M | 5412 | hCoV-19/Scotland/QEUH-9E3635/2020\|EPI_ISL_585510\|2 | .................... | .................... | .................... | .................... | .................... | .................... |
| M | 5413 | hCoV-19/Scotland/QEUH-9E3644/2020\|EPI_ISL_585511\|2 | .................... | .................... | .................... | .................... | .................... | .................... |
| M | 5414 | hCoV-19/Scotland/QEUH-9E3820/2020\|EPI_ISL_585515\|2 | .................... | .................... | .................... | .................... | .................... | .................... |
| M | 5415 | hCoV-19/Scotland/QEUH-9E383F/2020\|EPI_ISL_585516\|2 | .................... | .................... | .................... | .................... | .................... | .................... |
| M | 5416 | hCoV-19/Scotland/QEUH-9E384E/2020\|EPI_ISL_585517\|2 | .................... | .................... | .................... | .................... | .................... | .................... |
| M | 5417 | hCoV-19/Scotland/QEUH-9E386C/2020\|EPI_ISL_585519\|2 | .................... | .................... | .................... | .................... | .................... | .................... |
| M | 5418 | hCoV-19/Scotland/QEUH-9E38C6/2020\|EPI_ISL_585524\|2 | .................... | .A.......A.......... | .................... | .................... | .................... | .................... |
| M | 5419 | hCoV-19/Scotland/QEUH-9E393C/2020\|EPI_ISL_585529\|2 | .................... | .................... | .................... | .................... | .................... | .................... |
| M | 5420 | hCoV-19/Scotland/QEUH-9E39A5/2020\|EPI_ISL_585531\|2 | .................... | .A.................. | .................... | .................... | .................... | .................... |
| M | 5421 | hCoV-19/Scotland/QEUH-A005BC/2020\|EPI_ISL_585533\|2 | .................... | .................... | .................... | .................... | .................... | .................... |
| M | 5422 | hCoV-19/Scotland/QEUH-A009CF/2020\|EPI_ISL_585536\|2 | .................... | .................... | .................... | .................... | .................... | .................... |
| M | 5423 | hCoV-19/Scotland/QEUH-A009DE/2020\|EPI_ISL_585537\|2 | .................... | .................... | .................... | .................... | .................... | .................... |
| M | 5424 | hCoV-19/Scotland/QEUH-A009FC/2020\|EPI_ISL_585538\|2 | .................... | .................... | .................... | .................... | .................... | .................... |
| M | 5425 | hCoV-19/Scotland/QEUH-A00A26/2020\|EPI_ISL_585540\|2 | .................... | .................... | .................... | .................... | .................... | .................... |
| M | 5426 | hCoV-19/Scotland/QEUH-A00A35/2020\|EPI_ISL_585541\|2 | .................... | .................... | .................... | .................... | ......T............. | .................... |
| M | 5428 | hCoV-19/Scotland/QEUH-A00A71/2020\|EPI_ISL_585543\|2 | .................... | .................... | .................... | .................... | .................... | .................... |
| M | 5429 | hCoV-19/Scotland/QEUH-A00ACC/2020\|EPI_ISL_585546\|2 | .................... | .................... | .................... | .................... | .................... | .................... |
| M | 5432 | hCoV-19/Scotland/QEUH-A00B23/2020\|EPI_ISL_585550\|2 | .................... | .................... | .................... | .................... | .................... | .................... |
| M | 5433 | hCoV-19/Scotland/QEUH-A00B50/2020\|EPI_ISL_585551\|2 | .................... | .................... | .................... | .................... | .................... | .................... |
| M | 5434 | hCoV-19/Scotland/QEUH-A00B6F/2020\|EPI_ISL_585552\|2 | .................... | .................... | .................... | .................... | .................... | .................... |
| M | 5436 | hCoV-19/Scotland/QEUH-A00BAB/2020\|EPI_ISL_585556\|2 | .................... | .................... | .................... | .................... | .................... | .................... |
| M | 5437 | hCoV-19/Scotland/QEUH-A00BBA/2020\|EPI_ISL_585557\|2 | .................... | .................... | .................... | .................... | .................... | .................... |
| M | 5438 | hCoV-19/Scotland/QEUH-A00BD8/2020\|EPI_ISL_585558\|2 | .................... | .................... | .................... | .................... | .................... | .................... |
| M | 5439 | hCoV-19/Scotland/QEUH-A00BE7/2020\|EPI_ISL_585559\|2 | .................... | .................... | .................... | .................... | .................... | .................... |
| M | 5440 | hCoV-19/Scotland/QEUH-A00C02/2020\|EPI_ISL_585560\|2 | .................... | .................... | .................... | .................... | .................... | .................... |
| M | 5441 | hCoV-19/Scotland/QEUH-A00C11/2020\|EPI_ISL_585561\|2 | .................... | .................... | .................... | .................... | .................... | .................... |
| M | 5442 | hCoV-19/Scotland/QEUH-A00C3F/2020\|EPI_ISL_585562\|2 | .................... | .................... | .................... | .................... | .................... | .................... |
| M | 5443 | hCoV-19/Scotland/QEUH-A00C7B/2020\|EPI_ISL_585565\|2 | .................... | .................... | .................... | .................... | .................... | .................... |
| M | 5444 | hCoV-19/Scotland/QEUH-A00C99/2020\|EPI_ISL_585567\|2 | .................... | .................... | .................... | .................... | .................... | .................... |
| M | 5446 | hCoV-19/Scotland/QEUH-A00CD5/2020\|EPI_ISL_585570\|2 | .................... | .................... | .................... | .................... | .................... | .................... |
| M | 5447 | hCoV-19/Scotland/QEUH-A00D1E/2020\|EPI_ISL_585571\|2 | .................... | .................... | .................... | .................... | .................... | .................... |
| M | 5448 | hCoV-19/Scotland/QEUH-A00DB4/2020\|EPI_ISL_585573\|2 | .................... | .................... | .................... | .................... | .................... | .................... |
| M | 5449 | hCoV-19/Scotland/QEUH-A00DC3/2020\|EPI_ISL_585574\|2 | .................... | .................... | .................... | .................... | .................... | .................... |
| M | 5450 | hCoV-19/Scotland/QEUH-A00E1B/2020\|EPI_ISL_585578\|2 | .................... | .................... | .................... | .................... | .................... | .................... |
| M | 5451 | hCoV-19/Scotland/QEUH-A0113F/2020\|EPI_ISL_585580\|2 | .................... | .................... | .................... | .................... | .................... | .................... |
| M | 5452 | hCoV-19/Scotland/QEUH-A0117B/2020\|EPI_ISL_585581\|2 | .................... | .................... | .................... | .................... | .................... | .................... |

| | | | | | | | | |
|---|---|---|---|---|---|---|---|---|
| M | 5453 | hCoV-19/Scotland/QEUH-A011A8/2020\|EPI_ISL_585582\|2 | ···················· | ···················· | ···················· | ···················· | ···················· | ···················· |
| M | 5455 | hCoV-19/England/ALDP-A2DC87/2020\|EPI_ISL_586820\|20 | ···················· | ···················· | ···················· | ···················· | ···················· | ···················· |
| M | 5456 | hCoV-19/England/ALDP-A2D2D0/2020\|EPI_ISL_586821\|20 | ···················· | ···················· | ···················· | ···················· | ···················· | ···················· |
| M | 5457 | hCoV-19/England/ALDP-A2DA14/2020\|EPI_ISL_586822\|20 | ···················· | ···················· | ···················· | ···················· | ···················· | ···················· |
| M | 5458 | hCoV-19/England/ALDP-A2DBD5/2020\|EPI_ISL_586823\|20 | ···················· | ···················· | ···················· | ···················· | ···················· | ···················· |
| M | 5459 | hCoV-19/England/ALDP-A2D980/2020\|EPI_ISL_586824\|20 | ···················· | ···················· | ···················· | ···················· | ···················· | ···················· |
| M | 5460 | hCoV-19/England/ALDP-A2E2B1/2020\|EPI_ISL_586825\|20 | ···················· | ···················· | ···················· | ···················· | ···················· | ···················· |
| M | 5461 | hCoV-19/England/ALDP-A2DF15/2020\|EPI_ISL_586826\|20 | ···················· | ···················· | ···················· | ···················· | ···················· | ···················· |
| M | 5462 | hCoV-19/England/ALDP-A2E169/2020\|EPI_ISL_586827\|20 | ···················· | ···················· | ···················· | ···················· | ···················· | ···················· |
| M | 5463 | hCoV-19/England/ALDP-A2D05E/2020\|EPI_ISL_586828\|20 | ···················· | ···················· | ···················· | ···················· | ···················· | ···················· |
| M | 5464 | hCoV-19/England/ALDP-A2DD93/2020\|EPI_ISL_586829\|20 | ···················· | ···················· | ···················· | ···················· | ···················· | ···················· |
| M | 5465 | hCoV-19/England/ALDP-A2E1C3/2020\|EPI_ISL_586830\|20 | ···················· | ···················· | ···················· | ···················· | ···················· | ···················· |
| M | 5466 | hCoV-19/England/ALDP-A2D883/2020\|EPI_ISL_586831\|20 | ···················· | ···················· | ···················· | ···················· | ···················· | ···················· |
| M | 5467 | hCoV-19/England/ALDP-A2E3EB/2020\|EPI_ISL_586832\|20 | ···················· | ···················· | ···················· | ···················· | ···················· | ···················· |
| M | 5468 | hCoV-19/England/ALDP-A2D08B/2020\|EPI_ISL_586833\|20 | ···················· | ···················· | ···················· | ···················· | ···················· | ···················· |
| M | 5469 | hCoV-19/England/ALDP-A2D1E2/2020\|EPI_ISL_586834\|20 | ···················· | ···················· | ···················· | ···················· | ···················· | ···················· |
| M | 5470 | hCoV-19/England/ALDP-A2D59B/2020\|EPI_ISL_586835\|20 | ···················· | ···················· | ···················· | ···················· | ···················· | ···················· |
| M | 5471 | hCoV-19/England/ALDP-A2D2C1/2020\|EPI_ISL_586836\|20 | ···················· | ···················· | ···················· | ···················· | ···················· | ···················· |
| M | 5472 | hCoV-19/England/ALDP-A2D5C8/2020\|EPI_ISL_586837\|20 | ···················· | ···················· | ···················· | ···················· | ···················· | ···················· |
| M | 5473 | hCoV-19/England/ALDP-A2D100/2020\|EPI_ISL_586838\|20 | ···················· | ···················· | ···················· | ···················· | ···················· | ···················· |
| M | 5474 | hCoV-19/England/ALDP-A2D346/2020\|EPI_ISL_586839\|20 | ···················· | ···················· | ···················· | ···················· | ···················· | ···················· |
| M | 5475 | hCoV-19/England/ALDP-A2D8B0/2020\|EPI_ISL_586840\|20 | ···················· | ···················· | ···················· | ···················· | ···················· | ···················· |
| M | 5476 | hCoV-19/England/ALDP-A2E4D9/2020\|EPI_ISL_586841\|20 | ···················· | ···················· | ···················· | ···················· | ···················· | ···················· |
| M | 5477 | hCoV-19/England/ALDP-A2DB7B/2020\|EPI_ISL_586842\|20 | ···················· | ·A·················· | ···················· | ···················· | ···················· | ···················· |
| M | 5478 | hCoV-19/England/ALDP-A2E2EE/2020\|EPI_ISL_586843\|20 | ···················· | ·A·················· | ···················· | ···················· | ···················· | ···················· |
| M | 5479 | hCoV-19/England/ALDP-A2D373/2020\|EPI_ISL_586844\|20 | ···················· | ···················· | ···················· | ···················· | ···················· | ···················· |
| M | 5480 | hCoV-19/England/ALDP-A2E424/2020\|EPI_ISL_586845\|20 | ···················· | ···················· | ···················· | ···················· | ···················· | ···················· |
| M | 5481 | hCoV-19/England/ALDP-A2E512/2020\|EPI_ISL_586846\|20 | ···················· | ···················· | ···················· | ···················· | ···················· | ···················· |
| M | 5482 | hCoV-19/England/ALDP-A2D892/2020\|EPI_ISL_586848\|20 | ···················· | ···················· | ···················· | ···················· | ···················· | ···················· |
| M | 5483 | hCoV-19/England/ALDP-A2E0D5/2020\|EPI_ISL_586849\|20 | ···················· | ···················· | ···················· | ···················· | ···················· | ···················· |
| M | 5484 | hCoV-19/England/ALDP-A2E257/2020\|EPI_ISL_586850\|20 | ···················· | ···················· | ···················· | ···················· | ···················· | ···················· |
| M | 5485 | hCoV-19/England/ALDP-A2DFD9/2020\|EPI_ISL_586851\|20 | ···················· | ···················· | ···················· | ···················· | ···················· | ···················· |
| M | 5486 | hCoV-19/England/ALDP-A2DD48/2020\|EPI_ISL_586852\|20 | ···················· | ···················· | ···················· | ···················· | ···················· | ···················· |
| M | 5487 | hCoV-19/England/ALDP-A2D7D1/2020\|EPI_ISL_586853\|20 | ···················· | ···················· | ···················· | ···················· | ···················· | ···················· |
| M | 5488 | hCoV-19/England/ALDP-A2D610/2020\|EPI_ISL_586854\|20 | ···················· | ···················· | ···················· | ···················· | ···················· | ···················· |
| M | 5489 | hCoV-19/England/ALDP-A2E503/2020\|EPI_ISL_586856\|20 | ···················· | ···················· | ···················· | ···················· | ···················· | ···················· |
| M | 5490 | hCoV-19/England/ALDP-A2DDFD/2020\|EPI_ISL_586858\|20 | ···················· | ···················· | ···················· | ···················· | ···················· | ···················· |
| M | 5491 | hCoV-19/England/ALDP-A2D944/2020\|EPI_ISL_586859\|20 | ···················· | ···················· | ···················· | ···················· | ···················· | ···················· |
| M | 5492 | hCoV-19/England/ALDP-A2E54F/2020\|EPI_ISL_586860\|20 | ···················· | ···················· | ···················· | ···················· | ···················· | ···················· |
| M | 5493 | hCoV-19/England/ALDP-A2E4AC/2020\|EPI_ISL_586861\|20 | ···················· | ···················· | ···················· | ···················· | ···················· | ···················· |
| M | 5494 | hCoV-19/England/ALDP-A2D2EF/2020\|EPI_ISL_586862\|20 | ···················· | ···················· | ···················· | ···················· | ···················· | ···················· |
| M | 5495 | hCoV-19/England/ALDP-A2DC4B/2020\|EPI_ISL_586863\|20 | ···················· | ···················· | ···················· | ···················· | ···················· | ···················· |
| M | 5496 | hCoV-19/England/ALDP-A2E61F/2020\|EPI_ISL_586864\|20 | ···················· | ···················· | ···················· | ···················· | ···················· | ···················· |
| M | 5497 | hCoV-19/England/ALDP-A2E4E8/2020\|EPI_ISL_586865\|20 | ···················· | ···················· | ···················· | ···················· | ···················· | ···················· |
| M | 5498 | hCoV-19/England/ALDP-A2D7E0/2020\|EPI_ISL_586866\|20 | ···················· | ···················· | ···················· | ···················· | ···················· | ···················· |
| M | 5499 | hCoV-19/England/ALDP-A2E767/2020\|EPI_ISL_586867\|20 | ···················· | ···················· | ···················· | ···················· | ···················· | ···················· |
| M | 5500 | hCoV-19/England/ALDP-A2DCF0/2020\|EPI_ISL_586868\|20 | ···················· | ···················· | ···················· | ···················· | ···················· | ···················· |
| M | 5501 | hCoV-19/England/ALDP-A2D8CF/2020\|EPI_ISL_586869\|20 | ···················· | ···················· | ···················· | ···················· | ···················· | ···················· |
| M | 5502 | hCoV-19/England/ALDP-A2D425/2020\|EPI_ISL_586870\|20 | ···················· | ···················· | ···················· | ···················· | ···················· | ···················· |
| M | 5503 | hCoV-19/England/ALDP-A2DD1B/2020\|EPI_ISL_586871\|20 | ···················· | ···················· | ···················· | ···················· | ···················· | ···················· |
| M | 5504 | hCoV-19/England/ALDP-A2E239/2020\|EPI_ISL_586872\|20 | ···················· | ···················· | ···················· | ···················· | ···················· | ···················· |
| M | 5505 | hCoV-19/England/ALDP-A2D65C/2020\|EPI_ISL_586873\|20 | ···················· | ···················· | ···················· | ···················· | ···················· | ···················· |
| M | 5506 | hCoV-19/England/ALDP-A2E0F3/2020\|EPI_ISL_586874\|20 | ···················· | ···················· | ···················· | ···················· | ···················· | ···················· |
| M | 5507 | hCoV-19/England/ALDP-A2D197/2020\|EPI_ISL_586876\|20 | ···················· | ···················· | ···················· | ···················· | ···················· | ···················· |
| M | 5508 | hCoV-19/England/ALDP-A2D15B/2020\|EPI_ISL_586877\|20 | ···················· | ···················· | ···················· | ···················· | ···················· | ···················· |
| M | 5509 | hCoV-19/England/ALDP-A2E442/2020\|EPI_ISL_586878\|20 | ···················· | ···················· | ···················· | ···················· | ···················· | ···················· |
| M | 5510 | hCoV-19/England/ALDP-A2DCE1/2020\|EPI_ISL_586879\|20 | ···················· | ···················· | ···················· | ···················· | ···················· | ···················· |
| M | 5511 | hCoV-19/England/ALDP-A2D249/2020\|EPI_ISL_586880\|20 | ···················· | ···················· | ···················· | ···················· | ···················· | ···················· |
| M | 5512 | hCoV-19/England/ALDP-A2E5C7/2020\|EPI_ISL_586881\|20 | ···················· | ···················· | ···················· | ···················· | ···················· | ···················· |
| M | 5513 | hCoV-19/England/ALDP-A2D6E3/2020\|EPI_ISL_586882\|20 | ···················· | ···················· | ···················· | ···················· | ···················· | ···················· |
| M | 5514 | hCoV-19/England/ALDP-A2D1C4/2020\|EPI_ISL_586883\|20 | ···················· | ···················· | ···················· | ···················· | ···················· | ···················· |
| M | 5515 | hCoV-19/England/ALDP-A2D829/2020\|EPI_ISL_586885\|20 | ···················· | ···················· | ···················· | ···················· | ···················· | ···················· |
| M | 5516 | hCoV-19/England/ALDP-A2D504/2020\|EPI_ISL_586886\|20 | ···················· | ···················· | ···················· | ···················· | ···················· | ···················· |
| M | 5517 | hCoV-19/England/ALDP-A2D0D6/2020\|EPI_ISL_586888\|20 | ···················· | ···················· | ···················· | ···················· | ···················· | ···················· |
| M | 5518 | hCoV-19/England/ALDP-A2E178/2020\|EPI_ISL_586889\|20 | ···················· | ···················· | ···················· | ···················· | ···················· | ···················· |
| M | 5519 | hCoV-19/England/ALDP-A2D258/2020\|EPI_ISL_586890\|20 | ···················· | ···················· | ···················· | ···················· | ···················· | ···················· |
| M | 5520 | hCoV-19/England/ALDP-A2DDEE/2020\|EPI_ISL_586891\|20 | ···················· | ···················· | ···················· | ···················· | ···················· | ···················· |
| M | 5521 | hCoV-19/England/ALDP-A2D04F/2020\|EPI_ISL_586892\|20 | ···················· | ···················· | ···················· | ···················· | ···················· | ···················· |
| M | 5522 | hCoV-19/England/ALDP-A2E415/2020\|EPI_ISL_586893\|20 | ···················· | ···················· | ···················· | ···················· | ···················· | ···················· |
| M | 5523 | hCoV-19/England/ALDP-A2E0E4/2020\|EPI_ISL_586894\|20 | ···················· | ···················· | ···················· | ···················· | ···················· | ···················· |
| M | 5524 | hCoV-19/England/ALDP-A2E1B4/2020\|EPI_ISL_586895\|20 | ···················· | ···················· | ···················· | ···················· | ···················· | ···················· |
| M | 5525 | hCoV-19/England/ALDP-A2E10F/2020\|EPI_ISL_586896\|20 | ···················· | ···················· | ···················· | ···················· | ···················· | ···················· |
| M | 5526 | hCoV-19/England/ALDP-A2D337/2020\|EPI_ISL_586897\|20 | ···················· | ···················· | ···················· | ···················· | ···················· | ···················· |
| M | 5527 | hCoV-19/England/ALDP-A2D443/2020\|EPI_ISL_586898\|20 | ···················· | ···················· | ···················· | ···················· | ···················· | ···················· |
| M | 5528 | hCoV-19/England/ALDP-A2E248/2020\|EPI_ISL_586899\|20 | ···················· | ···················· | ···················· | ···················· | ···················· | ···················· |
| M | 5529 | hCoV-19/England/QEUH-9CF09B/2020\|EPI_ISL_590086\|20 | ···················· | ···················· | ···················· | ···················· | ···················· | ···················· |
| M | 5530 | hCoV-19/England/QEUH-9CF70F/2020\|EPI_ISL_590087\|20 | ···················· | ···················· | ···················· | ···················· | ···················· | ···················· |
| M | 5531 | hCoV-19/England/QEUH-9CEC98/2020\|EPI_ISL_590088\|20 | ···················· | ···················· | ···················· | ···················· | ···················· | ···················· |
| M | 5532 | hCoV-19/England/QEUH-9CEA43/2020\|EPI_ISL_590089\|20 | ···················· | ···················· | ···················· | ···················· | ···················· | ···················· |
| M | 5533 | hCoV-19/England/QEUH-9CED95/2020\|EPI_ISL_590090\|20 | ···················· | ···················· | ···················· | ···················· | ···················· | ···················· |

| | | | | | | | | |
|---|---|---|---|---|---|---|---|---|
| M | 5534 | hCoV-19/England/QEUH-9CF24A/2020\|EPI_ISL_590091\|20 | ···················· | ···················· | ···················· | ···················· | ···················· | ···················· |
| M | 5535 | hCoV-19/England/QEUH-9CE81C/2020\|EPI_ISL_590092\|20 | ···················· | ···················· | ···················· | ···················· | ···················· | ···················· |
| M | 5536 | hCoV-19/England/QEUH-9CF602/2020\|EPI_ISL_590093\|20 | ···················· | ·A·················· | ···················· | ···················· | ···················· | ···················· |
| M | 5537 | hCoV-19/England/QEUH-9CF4EA/2020\|EPI_ISL_590094\|20 | ···················· | ···················· | ···················· | ···················· | ···················· | ···················· |
| M | 5538 | hCoV-19/England/QEUH-9CF3DE/2020\|EPI_ISL_590095\|20 | ···················· | ···················· | ···················· | ···················· | ···················· | ···················· |
| M | 5539 | hCoV-19/England/QEUH-9CE894/2020\|EPI_ISL_590096\|20 | ···················· | ·····C·············· | ···················· | ···················· | ···················· | ···················· |
| M | 5540 | hCoV-19/England/QEUH-9CE1E4/2020\|EPI_ISL_590097\|20 | ···················· | ···················· | ···················· | ···················· | ···················· | ···················· |
| M | 5541 | hCoV-19/England/QEUH-9CE8D0/2020\|EPI_ISL_590098\|20 | ···················· | ···················· | ···················· | ···················· | ···················· | ···················· |
| M | 5542 | hCoV-19/England/QEUH-9CE7E2/2020\|EPI_ISL_590099\|20 | ···················· | ···················· | ···················· | ···················· | ···················· | ···················· |
| M | 5543 | hCoV-19/Canada/ON-S793/2020\|EPI_ISL_591230\|2020-04 | ···················· | ···················· | ···················· | ···················· | ···················· | ···················· |
| M | 5544 | hCoV-19/Canada/ON-S796/2020\|EPI_ISL_591231\|2020-04 | ···················· | ···················· | ···················· | ···················· | ···················· | ···················· |
| M | 5545 | hCoV-19/Canada/ON-S798/2020\|EPI_ISL_591232\|2020-04 | ···················· | ···················· | ···················· | ···················· | ···················· | ···················· |
| M | 5546 | hCoV-19/Canada/ON-S799/2020\|EPI_ISL_591233\|2020-04 | ···················· | ···················· | ···················· | ···················· | ···················· | ···················· |
| M | 5547 | hCoV-19/Canada/ON-S805/2020\|EPI_ISL_591234\|2020-04 | ···················· | ···················· | ···················· | ···················· | ···················· | ···················· |
| M | 5548 | hCoV-19/Canada/ON-S806/2020\|EPI_ISL_591235\|2020-04 | ···················· | ···················· | ···················· | ···················· | ···················· | ···················· |
| M | 5549 | hCoV-19/Canada/ON-S808/2020\|EPI_ISL_591236\|2020-04 | ···················· | ···················· | ···················· | ···················· | ···················· | ···················· |
| M | 5550 | hCoV-19/Canada/ON-S810/2020\|EPI_ISL_591237\|2020-04 | ···················· | ···················· | ···················· | ···················· | ···················· | ···················· |
| M | 5551 | hCoV-19/Canada/ON-S812/2020\|EPI_ISL_591238\|2020-04 | ···················· | ···················· | ···················· | ···················· | ···················· | ···················· |
| M | 5552 | hCoV-19/Canada/ON-S821/2020\|EPI_ISL_591239\|2020-04 | ···················· | ···················· | ···················· | ···················· | ···················· | ···················· |
| M | 5553 | hCoV-19/Canada/ON-S826/2020\|EPI_ISL_591240\|2020-04 | ···················· | ···················· | ···················· | ···················· | ···················· | ···················· |
| M | 5554 | hCoV-19/Canada/ON-S836/2020\|EPI_ISL_591241\|2020-04 | ···················· | ···················· | ···················· | ···················· | ···················· | ···················· |
| M | 5555 | hCoV-19/Canada/ON-S839/2020\|EPI_ISL_591242\|2020-04 | ···················· | ···················· | ···················· | ···················· | ···················· | ···················· |
| M | 5556 | hCoV-19/Canada/ON-S843/2020\|EPI_ISL_591244\|2020-04 | ···················· | ···················· | ···················· | ···················· | ···················· | ···················· |
| M | 5557 | hCoV-19/Canada/ON-S848/2020\|EPI_ISL_591245\|2020-04 | ···················· | ···················· | ···················· | ···················· | ···················· | ···················· |
| M | 5558 | hCoV-19/Canada/ON-S849/2020\|EPI_ISL_591246\|2020-04 | ···················· | ···················· | ···················· | ···················· | ···················· | ···················· |
| M | 5559 | hCoV-19/Canada/ON-S850/2020\|EPI_ISL_591247\|2020-04 | ···················· | ···················· | ···················· | ···················· | ···················· | ···················· |
| M | 5560 | hCoV-19/Canada/ON-S862/2020\|EPI_ISL_591248\|2020-04 | ···················· | ···················· | ···················· | ···················· | ···················· | ···················· |
| M | 5561 | hCoV-19/Canada/ON-S863/2020\|EPI_ISL_591249\|2020-04 | ···················· | ···················· | ···················· | ···················· | ···················· | ···················· |
| M | 5562 | hCoV-19/Canada/ON-S868/2020\|EPI_ISL_591250\|2020-04 | ···················· | ···················· | ···················· | ···················· | ···················· | ···················· |
| M | 5563 | hCoV-19/France/OCC-61/2020\|EPI_ISL_593876\|2020-07- | ···················· | ···················· | ···················· | ···················· | ···················· | ···················· |
| M | 5564 | hCoV-19/France/OCC-65/2020\|EPI_ISL_593880\|2020-08- | ···················· | ···················· | ···················· | ···················· | ···················· | ···················· |
| M | 5565 | hCoV-19/France/OCC-81/2020\|EPI_ISL_593892\|2020-06- | ···················· | ···················· | ···················· | ···················· | ···················· | ···················· |
| M | 5566 | hCoV-19/France/OCC-85/2020\|EPI_ISL_593893\|2020-09- | ···················· | ···················· | ···················· | ···················· | ···················· | ···················· |
| M | 5567 | hCoV-19/France/OCC-86/2020\|EPI_ISL_593894\|2020-09- | ···················· | ···················· | ···················· | ···················· | ···················· | ···················· |
| M | 5568 | hCoV-19/France/OCC-87/2020\|EPI_ISL_593895\|2020-10- | ···················· | ···················· | ···················· | ···················· | ···················· | ···················· |
| M | 5569 | hCoV-19/France/OCC-88/2020\|EPI_ISL_593896\|2020-10- | ···················· | ···················· | ···················· | ···················· | ···················· | ···················· |
| M | 5570 | hCoV-19/France/OCC-89/2020\|EPI_ISL_593897\|2020-10- | ···················· | ···················· | ···················· | ···················· | ···················· | ···················· |
| M | 5571 | hCoV-19/France/OCC-90/2020\|EPI_ISL_593898\|2020-10- | ···················· | ···················· | ···················· | ···················· | ···················· | ···················· |
| M | 5572 | hCoV-19/France/OCC-91/2020\|EPI_ISL_593899\|2020-10- | ···················· | ···················· | ···················· | ···················· | ···················· | ···················· |
| M | 5573 | hCoV-19/England/SHEF-C7551/2020\|EPI_ISL_596202\|202 | ···················· | ···················· | ···················· | ···················· | ···················· | ···················· |
| M | 5574 | hCoV-19/England/SHEF-C8CA6/2020\|EPI_ISL_596205\|202 | ···················· | ···················· | ···················· | ···················· | ···················· | ···················· |
| M | 5575 | hCoV-19/England/SHEF-C9443/2020\|EPI_ISL_596206\|202 | ···················· | ···················· | ···················· | ···················· | ···················· | ···················· |
| M | 5576 | hCoV-19/England/SHEF-C971D/2020\|EPI_ISL_596207\|202 | ···················· | ···················· | ···················· | ···················· | ···················· | ···················· |
| M | 5577 | hCoV-19/England/SHEF-CA837/2020\|EPI_ISL_596209\|202 | ···················· | ···················· | ···················· | ···················· | ···················· | ···················· |
| M | 5578 | hCoV-19/England/SHEF-CABA7/2020\|EPI_ISL_596210\|202 | ···················· | ·A·················· | ···················· | ···················· | ···················· | ···················· |
| M | 5579 | hCoV-19/England/SHEF-CD4C7/2020\|EPI_ISL_596213\|202 | ···················· | ···················· | ···················· | ···················· | ···················· | ···················· |
| M | 5580 | hCoV-19/England/SHEF-CDE9C/2020\|EPI_ISL_596215\|202 | ···················· | ···················· | ···················· | ···················· | ···················· | ···················· |
| M | 5581 | hCoV-19/England/SHEF-CE569/2020\|EPI_ISL_596216\|202 | ···················· | ·A·················· | ···················· | ···················· | ···················· | ···················· |
| M | 5582 | hCoV-19/England/SHEF-CF6B0/2020\|EPI_ISL_596218\|202 | ···················· | ···················· | ···················· | ···················· | ···················· | ···················· |
| M | 5583 | hCoV-19/England/SHEF-CFD33/2020\|EPI_ISL_596220\|202 | ···················· | ···················· | ···················· | ···················· | ···················· | ···················· |
| M | 5584 | hCoV-19/England/SHEF-D0995/2020\|EPI_ISL_596221\|202 | ···················· | ···················· | ···················· | ···················· | ···················· | ···················· |
| M | 5585 | hCoV-19/England/SHEF-D09FF/2020\|EPI_ISL_596222\|202 | ···················· | ···················· | ···················· | ···················· | ···················· | ···················· |
| M | 5586 | hCoV-19/England/SHEF-D0E0F/2020\|EPI_ISL_596223\|202 | ···················· | ···················· | ···················· | ···················· | ···················· | ···················· |
| M | 5587 | hCoV-19/England/SHEF-D18A6/2020\|EPI_ISL_596224\|202 | ···················· | ···················· | ···················· | ···················· | ···················· | ···················· |
| M | 5588 | hCoV-19/England/SHEF-D2016/2020\|EPI_ISL_596225\|202 | ···················· | ···················· | ···················· | ···················· | ···················· | ···················· |
| M | 5589 | hCoV-19/Russia/SPE-RII-30848S/2020\|EPI_ISL_596229\| | ···················· | ···················· | ···················· | ···················· | ···················· | ···················· |
| M | 5590 | hCoV-19/Russia/SPE-RII-32307S/2020\|EPI_ISL_596230\| | ···················· | ···················· | ···················· | ···················· | ···················· | ···················· |
| M | 5591 | hCoV-19/Russia/SPE-RII-32995S/2020\|EPI_ISL_596231\| | ···················· | ···················· | ···················· | ···················· | ···················· | ···················· |
| M | 5592 | hCoV-19/Russia/SPE-RII-34611S/2020\|EPI_ISL_596235\| | ···················· | ···················· | ···················· | ···················· | ···················· | ···················· |
| M | 5593 | hCoV-19/Russia/SPE-RII-34613S/2020\|EPI_ISL_596236\| | ···················· | ···················· | ···················· | ···················· | ···················· | ···················· |
| M | 5594 | hCoV-19/Russia/SPE-RII-34646S/2020\|EPI_ISL_596237\| | ···················· | ···················· | ···················· | ···················· | ···················· | ···················· |
| M | 5595 | hCoV-19/Russia/SPE-RII-34660S/2020\|EPI_ISL_596238\| | ···················· | ···················· | ···················· | ···················· | ···················· | ···················· |
| M | 5596 | hCoV-19/Russia/SPE-RII-34755S/2020\|EPI_ISL_596239\| | ···················· | ···················· | ···················· | ···················· | ···················· | ···················· |
| M | 5597 | hCoV-19/Russia/SPE-RII-34948S/2020\|EPI_ISL_596240\| | ···················· | ···················· | ···················· | ···················· | ···················· | ···················· |
| M | 5598 | hCoV-19/Russia/SPE-RII-35034S/2020\|EPI_ISL_596241\| | ···················· | ···················· | ···················· | ···················· | ···················· | ···················· |
| M | 5599 | hCoV-19/Russia/SPE-RII-35144S/2020\|EPI_ISL_596242\| | ···················· | ···················· | ···················· | ···················· | ···················· | ···················· |
| M | 5600 | hCoV-19/Russia/SPE-RII-35177S/2020\|EPI_ISL_596243\| | ···················· | ···················· | ···················· | ···················· | ···················· | ···················· |
| M | 5601 | hCoV-19/Russia/SPE-RII-35178S/2020\|EPI_ISL_596244\| | ···················· | ···················· | ···················· | ···················· | ···················· | ···················· |
| M | 5602 | hCoV-19/Russia/SPE-RII-35212S/2020\|EPI_ISL_596245\| | ···················· | ···················· | ···················· | ···················· | ···················· | ···················· |
| M | 5603 | hCoV-19/Russia/SPE-RII-MH1400S/2020\|EPI_ISL_596268 | ···················· | ···················· | ···················· | ···················· | ···················· | ···················· |
| M | 5604 | hCoV-19/Russia/NGR-RII-MH1404S/2020\|EPI_ISL_596269 | ···················· | ···················· | ···················· | ···················· | ···················· | ···················· |
| M | 5605 | hCoV-19/Russia/SVE-RII-MH1411S/2020\|EPI_ISL_596270 | ···················· | ···················· | ···················· | ···················· | ···················· | ···················· |
| M | 5606 | hCoV-19/Russia/SVE-RII-MH1412S/2020\|EPI_ISL_596271 | ···················· | ···················· | ···················· | ···················· | ···················· | ···················· |
| M | 5607 | hCoV-19/Russia/SVE-RII-MH1430S/2020\|EPI_ISL_596272 | ···················· | ···················· | ···················· | ···················· | ···················· | ···················· |
| M | 5608 | hCoV-19/Russia/SVE-RII-MH1433S/2020\|EPI_ISL_596273 | ···················· | ···················· | ···················· | ···················· | ···················· | ···················· |
| M | 5609 | hCoV-19/Russia/SVE-RII-MH1436S/2020\|EPI_ISL_596274 | ···················· | ···················· | ···················· | ···················· | ···················· | ···················· |
| M | 5610 | hCoV-19/Russia/SVE-RII-MH1438S/2020\|EPI_ISL_596275 | ···················· | ···················· | ···················· | ···················· | ···················· | ···················· |
| M | 5611 | hCoV-19/Russia/SVE-RII-MH1441S/2020\|EPI_ISL_596276 | ···················· | ···················· | ···················· | ···················· | ···················· | ···················· |
| M | 5612 | hCoV-19/Russia/SVE-RII-MH1443S/2020\|EPI_ISL_596277 | ···················· | ···················· | ···················· | ···················· | ···················· | ···················· |
| M | 5613 | hCoV-19/Russia/SVE-RII-MH1444S/2020\|EPI_ISL_596278 | ················G··· | ···················· | ···················· | ···················· | ···················· | ···················· |

| | | | | | | | |
|---|---|---|---|---|---|---|---|
| M | 5614 hCoV-19/Russia/SVE-RII-MH1452S/2020\|EPI_ISL_596279 | ···················· | ···················· | ····················· | ···················· | ···················· | ···················· |
| M | 5615 hCoV-19/Russia/SPE-RII-MH1464S/2020\|EPI_ISL_596280 | ···················· | ···················· | ····················· | ···················· | ···················· | ···················· |
| M | 5616 hCoV-19/Russia/SPE-RII-MH1489S/2020\|EPI_ISL_596281 | ···················· | ···················· | ····················· | ···················· | ···················· | ···················· |
| M | 5617 hCoV-19/Russia/SPE-RII-MH1500S/2020\|EPI_ISL_596282 | ···················· | ···················· | ····················· | ···················· | ···················· | ···················· |
| M | 5618 hCoV-19/Russia/SPE-RII-MH1504S/2020\|EPI_ISL_596283 | ···················· | ···················· | ····················· | ···················· | ···················· | ···················· |
| M | 5619 hCoV-19/Russia/SPE-RII-MH1509S/2020\|EPI_ISL_596284 | ···················· | ···················· | ····················· | ···················· | ···················· | ···················· |
| M | 5620 hCoV-19/Russia/SPE-RII-MH1512S/2020\|EPI_ISL_596285 | ···················· | ···················· | ····················· | ···················· | ···················· | ···················· |
| M | 5621 hCoV-19/Russia/SPE-RII-MH1515S/2020\|EPI_ISL_596286 | ···················· | ···················· | ····················· | ···················· | ···················· | ···················· |
| M | 5622 hCoV-19/Russia/SPE-RII-MH1519S/2020\|EPI_ISL_596288 | ···················· | ···················· | ····················· | ···················· | ···················· | ···················· |
| M | 5623 hCoV-19/Russia/SPE-RII-MH1520S/2020\|EPI_ISL_596289 | ···················· | ···················· | ····················· | ···················· | ···················· | ···················· |
| M | 5624 hCoV-19/Russia/SPE-RII-MH1524S/2020\|EPI_ISL_596291 | ···················· | ···················· | ····················· | ···················· | ···················· | ···················· |
| M | 5625 hCoV-19/Russia/SPE-RII-MH1535S/2020\|EPI_ISL_596292 | ···················· | ···················· | ····················· | ···················· | ···················· | ···················· |
| M | 5626 hCoV-19/Russia/SPE-RII-MH1568S/2020\|EPI_ISL_596293 | ···················· | ···················· | ····················· | ···················· | ···················· | ···················· |
| M | 5627 hCoV-19/Russia/SPE-RII-MH1609S/2020\|EPI_ISL_596294 | ···················· | ···················· | ····················· | ···················· | ···················· | ···················· |
| M | 5628 hCoV-19/Russia/SVE-RII-MH1612S/2020\|EPI_ISL_596295 | ···················· | ···················· | ····················· | ···················· | ···················· | ···················· |
| M | 5629 hCoV-19/Russia/SVE-RII-MH1613S/2020\|EPI_ISL_596296 | ···················· | ···················· | ····················· | ···················· | ···················· | ···················· |
| M | 5630 hCoV-19/Russia/SPE-RII-MH1616S/2020\|EPI_ISL_596297 | ···················· | ···················· | ····················· | ···················· | ···················· | ···················· |
| M | 5631 hCoV-19/Russia/SVE-RII-MH1618S/2020\|EPI_ISL_596298 | ···················· | ···················· | ····················· | ···················· | ···················· | ···················· |
| M | 5632 hCoV-19/Russia/SVE-RII-MH1626S/2020\|EPI_ISL_596299 | ···················· | ···················· | ····················· | ···················· | ···················· | ···················· |
| M | 5633 hCoV-19/USA/TX-DSHS-0729/2020\|EPI_ISL_584131\|2020- | ···················· | ···················· | ····················· | ···················· | ···················· | ···················· |
| M | 5634 hCoV-19/USA/TX-DSHS-0754/2020\|EPI_ISL_584155\|2020- | ···················· | ···················· | ····················· | ···················· | ···················· | ···················· |
| M | 5635 hCoV-19/USA/TX-DSHS-0767/2020\|EPI_ISL_584164\|2020- | ···················· | ···················· | ····················· | ···················· | ···················· | ···················· |
| M | 5637 hCoV-19/England/ALDP-9CC359/2020\|EPI_ISL_584166\|20 | ···················· | ···················· | ····················· | ···················· | ···················· | ···················· |
| M | 5638 hCoV-19/England/ALDP-9E5CB9/2020\|EPI_ISL_584167\|20 | ···················· | ···················· | ····················· | ···················· | ···················· | ···················· |
| M | 5640 hCoV-19/Scotland/EDB7941/2020\|EPI_ISL_585456\|2020- | ···················· | ···················· | ····················· | ···················· | ······T············· | ···················· |
| M | 5641 hCoV-19/Scotland/EDB7963/2020\|EPI_ISL_585461\|2020- | ···················· | ···················· | ····················· | ···················· | ···················· | ···················· |
| M | 5642 hCoV-19/Scotland/EDB7983/2020\|EPI_ISL_585463\|2020- | ···················· | ···················· | ····················· | ···················· | ···················· | ···················· |
| M | 5643 hCoV-19/Scotland/EDB8091/2020\|EPI_ISL_585477\|2020- | ···················· | ···················· | ····················· | ···················· | ···················· | ···················· |
| M | 5644 hCoV-19/Scotland/EDB8092/2020\|EPI_ISL_585478\|2020- | ···················· | ···················· | ····················· | ···················· | ···················· | ···················· |
| M | 5645 hCoV-19/Scotland/EDB8112/2020\|EPI_ISL_585479\|2020- | ···················· | ·A·················· | ····················· | ···················· | ···················· | ···················· |
| M | 5646 hCoV-19/Scotland/EDB8119/2020\|EPI_ISL_585480\|2020- | ···················· | ···················· | ····················· | ···················· | ···················· | ···················· |
| M | 5647 hCoV-19/Scotland/EDB8140/2020\|EPI_ISL_585481\|2020- | ···················· | ···················· | ····················· | ···················· | ···················· | ···················· |
| M | 5648 hCoV-19/Scotland/EDB8145/2020\|EPI_ISL_585482\|2020- | ···················· | ···················· | ····················· | ···················· | ···················· | ···················· |
| M | 5649 hCoV-19/Scotland/EDB8146/2020\|EPI_ISL_585483\|2020- | ···················· | ···················· | ····················· | ···················· | ···················· | ···················· |
| M | 5650 hCoV-19/Scotland/EDB8160/2020\|EPI_ISL_585484\|2020- | ···················· | ···················· | ····················· | ···················· | ···················· | ···················· |
| M | 5651 hCoV-19/Scotland/EDB8216/2020\|EPI_ISL_585486\|2020- | ···················· | ···················· | ····················· | ···················· | ···················· | ···················· |
| M | 5652 hCoV-19/Scotland/EDB8229/2020\|EPI_ISL_585487\|2020- | ···················· | ···················· | ····················· | ···················· | ···················· | ···················· |
| M | 5653 hCoV-19/Scotland/EDB8241/2020\|EPI_ISL_585492\|2020- | ···················· | ···················· | ····················· | ···················· | ···················· | ···················· |
| M | 5654 hCoV-19/Scotland/EDB8243/2020\|EPI_ISL_585494\|2020- | ···················· | ···················· | ····················· | ···················· | ···················· | ···················· |
| M | 5655 hCoV-19/Scotland/EDB8244/2020\|EPI_ISL_585495\|2020- | ···················· | ···················· | ····················· | ···················· | ······T············· | ···················· |
| M | 5656 hCoV-19/Scotland/EDB8245/2020\|EPI_ISL_585496\|2020- | ···················· | ···················· | ····················· | ···················· | ···················· | ···················· |
| M | 5657 hCoV-19/Scotland/EDB8248/2020\|EPI_ISL_585498\|2020- | ···················· | ···················· | ····················· | ···················· | ···················· | ···················· |
| M | 5658 hCoV-19/England/ALDP-A2D2B2/2020\|EPI_ISL_586786\|20 | ···················· | ···················· | ····················· | ···················· | ···················· | ···················· |
| M | 5659 hCoV-19/England/ALDP-A2DB6C/2020\|EPI_ISL_586787\|20 | ···················· | ···················· | ····················· | ···················· | ···················· | ···················· |
| M | 5660 hCoV-19/England/ALDP-A2E3AF/2020\|EPI_ISL_586788\|20 | ···················· | ···················· | ····················· | ···················· | ···················· | ···················· |
| M | 5661 hCoV-19/England/ALDP-A2D470/2020\|EPI_ISL_586789\|20 | ···················· | ···················· | ····················· | ···················· | ···················· | ···················· |
| M | 5662 hCoV-19/England/ALDP-A2D434/2020\|EPI_ISL_586790\|20 | ···················· | ···················· | ····················· | ···················· | ···················· | ···················· |
| M | 5663 hCoV-19/England/ALDP-A2E59A/2020\|EPI_ISL_586791\|20 | ···················· | ···················· | ····················· | ···················· | ···················· | ···················· |
| M | 5664 hCoV-19/England/ALDP-A2D865/2020\|EPI_ISL_586792\|20 | ···················· | ···················· | ····················· | ···················· | ···················· | ···················· |
| M | 5665 hCoV-19/England/ALDP-A2D30A/2020\|EPI_ISL_586793\|20 | ···················· | ···················· | ····················· | ···················· | ···················· | ···················· |
| M | 5666 hCoV-19/England/ALDP-A2E73A/2020\|EPI_ISL_586794\|20 | ···················· | ···················· | ····················· | ···················· | ···················· | ···················· |
| M | 5667 hCoV-19/England/ALDP-A2D14C/2020\|EPI_ISL_586795\|20 | ···················· | ···················· | ····················· | ···················· | ···················· | ···················· |
| M | 5668 hCoV-19/England/ALDP-A2D3BF/2020\|EPI_ISL_586796\|20 | ···················· | ···················· | ····················· | ···················· | ···················· | ···················· |
| M | 5669 hCoV-19/England/ALDP-A2D09A/2020\|EPI_ISL_586798\|20 | ···················· | ···················· | ····················· | ···················· | ···················· | ···················· |
| M | 5670 hCoV-19/Canada/ON-S1439/2020\|EPI_ISL_591201\|2020-0 | ···················· | ···················· | ····················· | ···················· | ···················· | ···················· |
| M | 5671 hCoV-19/Canada/ON-S1440/2020\|EPI_ISL_591202\|2020-0 | ···················· | ···················· | ····················· | ···················· | ···················· | ···················· |
| M | 5672 hCoV-19/Canada/ON-S1444/2020\|EPI_ISL_591204\|2020-0 | ···················· | ···················· | ····················· | ···················· | ···················· | ···················· |
| M | 5673 hCoV-19/Canada/ON-S1446/2020\|EPI_ISL_591205\|2020-0 | ···················· | ·A·················· | ····················· | ···················· | ···················· | ···················· |
| M | 5674 hCoV-19/Canada/ON-S1456/2020\|EPI_ISL_591207\|2020-0 | ···················· | ·A·················· | ····················· | ···················· | ···················· | ···A················ |
| M | 5675 hCoV-19/Canada/ON-S737/2020\|EPI_ISL_591210\|2020-04 | ···················· | ···················· | ····················· | ···················· | ···················· | ···················· |
| M | 5676 hCoV-19/Canada/ON-S743/2020\|EPI_ISL_591211\|2020-04 | ···················· | ···················· | ····················· | ···················· | ···················· | ···A················ |
| M | 5677 hCoV-19/Canada/ON-S746/2020\|EPI_ISL_591212\|2020-04 | ···················· | ···················· | ····················· | ···················· | ···················· | ···················· |
| M | 5678 hCoV-19/Canada/ON-S747/2020\|EPI_ISL_591213\|2020-04 | ···················· | ···················· | ····················· | ···················· | ···················· | ···················· |
| M | 5679 hCoV-19/Canada/ON-S751/2020\|EPI_ISL_591214\|2020-04 | ···················· | ···················· | ····················· | ···················· | ···················· | ···················· |
| M | 5680 hCoV-19/Canada/ON-S752/2020\|EPI_ISL_591215\|2020-04 | ···················· | ···················· | ····················· | ···················· | ···················· | ···················· |
| M | 5681 hCoV-19/Canada/ON-S753/2020\|EPI_ISL_591216\|2020-04 | ···················· | ···················· | ····················· | ···················· | ···················· | ···················· |
| M | 5682 hCoV-19/Canada/ON-S756/2020\|EPI_ISL_591217\|2020-04 | ···················· | ···················· | ····················· | ···················· | ···················· | ···················· |
| M | 5683 hCoV-19/Canada/ON-S760/2020\|EPI_ISL_591218\|2020-04 | ···················· | ···················· | ····················· | ···················· | ···················· | ···················· |
| M | 5684 hCoV-19/Canada/ON-S762/2020\|EPI_ISL_591219\|2020-04 | ···················· | ···················· | ····················· | ···················· | ···················· | ···················· |
| M | 5685 hCoV-19/Canada/ON-S764/2020\|EPI_ISL_591220\|2020-04 | ···················· | ···················· | ····················· | ···················· | ···················· | ···················· |
| M | 5686 hCoV-19/Canada/ON-S767/2020\|EPI_ISL_591221\|2020-04 | ···················· | ···················· | ····················· | ···················· | ···················· | ···················· |
| M | 5687 hCoV-19/Canada/ON-S773/2020\|EPI_ISL_591222\|2020-04 | ···················· | ···················· | ····················· | ···················· | ···················· | ···················· |
| M | 5688 hCoV-19/Canada/ON-S775/2020\|EPI_ISL_591223\|2020-04 | ···················· | ···················· | ····················· | ···················· | ···················· | ···················· |
| M | 5689 hCoV-19/Canada/ON-S776/2020\|EPI_ISL_591224\|2020-04 | ···················· | ···················· | ····················· | ···················· | ···················· | ···················· |
| M | 5690 hCoV-19/Canada/ON-S778/2020\|EPI_ISL_591225\|2020-04 | ···················· | ···················· | ····················· | ···················· | ···················· | ···················· |
| M | 5691 hCoV-19/Canada/ON-S780/2020\|EPI_ISL_591226\|2020-04 | ···················· | ···················· | ····················· | ···················· | ···················· | ···················· |
| M | 5692 hCoV-19/Canada/ON-S786/2020\|EPI_ISL_591227\|2020-04 | ···················· | ···················· | ····················· | ···················· | ···················· | ···················· |
| M | 5693 hCoV-19/Canada/ON-S787/2020\|EPI_ISL_591228\|2020-04 | ···················· | ···················· | ····················· | ···················· | ········Y··········· | ···················· |
| M | 5694 hCoV-19/Canada/ON-S789/2020\|EPI_ISL_591229\|2020-04 | ···················· | ···················· | ····················· | ···················· | ···················· | ···················· |
| M | 5695 hCoV-19/Canada/ON-S881/2020\|EPI_ISL_591253\|2020-04 | ···················· | ···················· | ····················· | ···················· | ···················· | ···················· |

| | | | | | | | | | |
|---|---|---|---|---|---|---|---|---|---|
| M | 5696 | hCoV-19/Canada/ON-S883/2020\|EPI_ISL_591254\|2020-04 | ···················· | ···················· | ····················· | ···················· | ···················· | ···················· | |
| M | 5697 | hCoV-19/Canada/ON-S888/2020\|EPI_ISL_591255\|2020-04 | ···················· | ···················· | ····················· | ···················· | ···················· | ···················· | |
| M | 5698 | hCoV-19/Canada/ON-S894/2020\|EPI_ISL_591256\|2020-04 | ···················· | ···················· | ····················· | ···················· | ···················· | ···················· | |
| M | 5699 | hCoV-19/Canada/ON-S908/2020\|EPI_ISL_591258\|2020-04 | ···················· | ···················· | ····················· | ···················· | ···················· | ···················· | |
| M | 5700 | hCoV-19/Canada/ON-S910/2020\|EPI_ISL_591259\|2020-04 | ···················· | ···················· | ····················· | ···················· | ···················· | ···················· | |
| M | 5701 | hCoV-19/Canada/ON-S926/2020\|EPI_ISL_591260\|2020-04 | ···················· | ···················· | ····················· | ···················· | ···················· | ···················· | |
| M | 5702 | hCoV-19/Canada/ON-S927/2020\|EPI_ISL_591261\|2020-04 | ···················· | ···················· | ····················· | ···················· | ···················· | ···················· | |
| M | 5703 | hCoV-19/Canada/ON-S931/2020\|EPI_ISL_591263\|2020-04 | ···················· | ···················· | ····················· | ···················· | ···················· | ···················· | |
| M | 5704 | hCoV-19/Canada/ON-S942/2020\|EPI_ISL_591264\|2020-04 | ···················· | ···················· | ····················· | ···················· | ···················· | ···················· | |
| M | 5705 | hCoV-19/Canada/ON-S943/2020\|EPI_ISL_591265\|2020-04 | ···················· | ···················· | ····················· | ···················· | ···················· | ···················· | |
| M | 5706 | hCoV-19/Canada/ON-S944/2020\|EPI_ISL_591266\|2020-04 | ···················· | ···················· | ····················· | ···················· | ···················· | ···················· | |
| M | 5707 | hCoV-19/Canada/ON-S950/2020\|EPI_ISL_591267\|2020-04 | ···················· | ···················· | ····················· | ···················· | ···················· | ···················· | |
| M | 5708 | hCoV-19/Canada/ON-S951/2020\|EPI_ISL_591268\|2020-04 | ···················· | ···················· | ····················· | ···················· | ···················· | ···················· | |
| M | 5709 | hCoV-19/Qingdao/IVDC-01-10/2020\|EPI_ISL_591270\|202 | ···················· | ···················· | ····················· | ···················· | ···················· | ···················· | |
| M | 5710 | hCoV-19/Qingdao/IVDC-02-10/2020\|EPI_ISL_591271\|202 | ···················· | ···················· | ····················· | ···················· | ···················· | ···················· | |
| M | 5711 | hCoV-19/env/Qingdao/IVDC-03-10/2020\|EPI_ISL_591272 | ···················· | ···················· | ····················· | ···················· | ···················· | ···················· | |
| M | 5712 | hCoV-19/env/Qingdao/IVDC-04-10/2020\|EPI_ISL_591273 | ···················· | ···················· | ····················· | ···················· | ···················· | ···················· | |
| M | 5715 | hCoV-19/env/Qingdao/IVDC-07-10/2020\|EPI_ISL_591276 | ···················· | ···················· | ····················· | ···················· | ···················· | ···················· | |
| M | 5716 | hCoV-19/env/Qingdao/IVDC-08-10/2020\|EPI_ISL_591277 | ···················· | ···················· | ····················· | ···················· | ···················· | ···················· | |
| M | 5717 | hCoV-19/env/Qingdao/IVDC-09-10/2020\|EPI_ISL_591278 | ···················· | ···················· | ····················· | ···················· | ···················· | ···················· | |
| M | 5718 | hCoV-19/env/Qingdao/IVDC-010-10/2020\|EPI_ISL_59127 | ···················· | ···················· | ····················· | ···················· | ···················· | ···················· | |
| M | 5719 | hCoV-19/env/Qingdao/IVDC-011-10/2020\|EPI_ISL_59128 | ···················· | ···················· | ····················· | ···················· | ···················· | ···················· | |
| M | 5720 | hCoV-19/USA/UT-UPHL-04230_2/2020\|EPI_ISL_591281\|20 | ···················· | ···················· | ····················· | ···················· | ···················· | ···················· | |
| M | 5721 | hCoV-19/USA/UT-UPHL-04235_2/2020\|EPI_ISL_591284\|20 | ···················· | ···················· | ····················· | ···················· | ···················· | ···················· | |
| M | 5722 | hCoV-19/USA/UT-UPHL-04236_2/2020\|EPI_ISL_591285\|20 | ···················· | ···················· | ····················· | ···················· | ···················· | ···················· | |
| M | 5723 | hCoV-19/USA/UT-UPHL-04262_2/2020\|EPI_ISL_591287\|20 | ···················· | ···················· | ····················· | ···················· | ···················· | ···················· | |
| M | 5724 | hCoV-19/USA/UT-UPHL-04265_2/2020\|EPI_ISL_591288\|20 | ···················· | ···················· | ····················· | ···················· | ···················· | ···················· | |
| M | 5725 | hCoV-19/USA/UT-UPHL-04282_2/2020\|EPI_ISL_591289\|20 | ···················· | ···················· | ····················· | ···················· | ···················· | ···················· | |
| M | 5726 | hCoV-19/USA/UT-UPHL-04289_2/2020\|EPI_ISL_591291\|20 | ···················· | ···················· | ····················· | ···················· | ···················· | ···················· | |
| M | 5727 | hCoV-19/USA/UT-UPHL-04290_2/2020\|EPI_ISL_591292\|20 | ···················· | ···················· | ····················· | ···················· | ···················· | ···················· | |
| M | 5729 | hCoV-19/Australia/VIC14436/2020\|EPI_ISL_592501\|202 | ···················· | ···················· | ··K·················· | ···················· | ··R················· | ···················· | |
| M | 5730 | hCoV-19/Australia/VIC14439/2020\|EPI_ISL_592502\|202 | ···················· | ···················· | ··K·················· | ···················· | ···················· | ···················· | |
| M | 5731 | hCoV-19/Australia/VIC14441/2020\|EPI_ISL_592503\|202 | ···················· | ···················· | ····················· | ···················· | ···················· | ···················· | |
| M | 5732 | hCoV-19/Australia/VIC14443/2020\|EPI_ISL_592504\|202 | ···················· | ···················· | ····················· | ···················· | ···················· | ···················· | |
| M | 5733 | hCoV-19/Australia/VIC14448/2020\|EPI_ISL_592505\|202 | ···················· | ···················· | ····················· | ···················· | ···················· | ···················· | |
| M | 5734 | hCoV-19/Australia/VIC14449/2020\|EPI_ISL_592506\|202 | ···················· | ···················· | ····················· | ···················· | ···················· | ···················· | |
| M | 5735 | hCoV-19/Australia/VIC14457/2020\|EPI_ISL_592507\|202 | ···················· | ···················· | ····················· | ···················· | ···················· | ···················· | |
| M | 5736 | hCoV-19/Australia/VIC14472/2020\|EPI_ISL_592509\|202 | ···················· | ···················· | ····················· | ···················· | ··················NN | ···················· | 1 |
| M | 5737 | hCoV-19/Australia/VIC14474/2020\|EPI_ISL_592510\|202 | ···················· | ···················· | ····················· | ···················· | ···················· | ···················· | |
| M | 5738 | hCoV-19/Australia/VIC14475/2020\|EPI_ISL_592511\|202 | ···················· | ···················· | ····················· | ···················· | ··K················· | ···················· | |
| M | 5740 | hCoV-19/Australia/VIC14480/2020\|EPI_ISL_592513\|202 | ···················· | ···················· | ··K·················· | ···················· | ···················· | ···················· | |
| M | 5741 | hCoV-19/Australia/VIC14483/2020\|EPI_ISL_592514\|202 | ···················· | ···················· | ····················· | ···················· | ···················· | ···················· | |
| M | 5742 | hCoV-19/Australia/VIC14484/2020\|EPI_ISL_592515\|202 | ···················· | ···················· | ··K·················· | ···················· | ···················· | ····N··············· | |
| M | 5743 | hCoV-19/Australia/VIC14488/2020\|EPI_ISL_592516\|202 | ···················· | ···················· | ····················· | ···················· | ···················· | ···················· | |
| M | 5744 | hCoV-19/Australia/VIC14490/2020\|EPI_ISL_592517\|202 | ···················· | ···················· | ····················· | ···················· | ···················· | ···················· | |
| M | 5747 | hCoV-19/Australia/VIC14493/2020\|EPI_ISL_592520\|202 | ···················· | ···················· | ····················· | ···················· | ···················· | ···················· | |
| M | 5748 | hCoV-19/Australia/VIC14494/2020\|EPI_ISL_592521\|202 | ···················· | ···················· | ····················· | ···················· | ···················· | ···················· | |
| M | 5750 | hCoV-19/Australia/VIC14499/2020\|EPI_ISL_592523\|202 | ···················· | ···················· | ····················· | ···················· | ···················· | ···················· | |
| M | 5751 | hCoV-19/Australia/VIC14500/2020\|EPI_ISL_592524\|202 | ···················· | ···················· | ····················· | ···················· | ···················· | ···················· | |
| M | 5753 | hCoV-19/Australia/VIC14502/2020\|EPI_ISL_592526\|202 | ···················· | ·····A·············· | ····················· | ···················· | ···················· | ···················· | |
| M | 5754 | hCoV-19/Australia/VIC14504/2020\|EPI_ISL_592527\|202 | ···················· | ···················· | ····················· | ···················· | ···················· | ···················· | |
| M | 5756 | hCoV-19/Australia/VIC14507/2020\|EPI_ISL_592529\|202 | ···················· | ···················· | ····················· | ···················· | ···················· | ···················· | |
| M | 5758 | hCoV-19/England/204221408/2020\|EPI_ISL_593800\|2020 | ···················· | ···················· | ····················· | ···················· | ···················· | ···················· | |
| M | 5759 | hCoV-19/England/204221410/2020\|EPI_ISL_593801\|2020 | ···················· | ···················· | ····················· | ···················· | ···················· | ···················· | |
| M | 5760 | hCoV-19/England/204221413/2020\|EPI_ISL_593803\|2020 | ···················· | ···················· | ····················· | ···················· | ···················· | ···················· | |
| M | 5761 | hCoV-19/England/204221414/2020\|EPI_ISL_593804\|2020 | ···················· | ···················· | ····················· | ···················· | ···················· | ···················· | |
| M | 5762 | hCoV-19/England/204221427/2020\|EPI_ISL_593806\|2020 | ···················· | ···················· | ····················· | ···················· | ···················· | ···················· | |
| M | 5763 | hCoV-19/England/204221431/2020\|EPI_ISL_593807\|2020 | ···················· | ···················· | ····················· | ···················· | ···················· | ···················· | |
| M | 5764 | hCoV-19/England/204221432/2020\|EPI_ISL_593808\|2020 | ···················· | ···················· | ····················· | ···················· | ···················· | ···················· | |
| M | 5765 | hCoV-19/England/204221433/2020\|EPI_ISL_593809\|2020 | ···················· | ···················· | ····················· | ···················· | ···················· | ···················· | |
| M | 5766 | hCoV-19/England/204221435/2020\|EPI_ISL_593810\|2020 | ···················· | ···················· | ····················· | ···················· | ···················· | ···················· | |
| M | 5767 | hCoV-19/England/204221437/2020\|EPI_ISL_593811\|2020 | ···················· | ···················· | ····················· | ···················· | ···················· | ···················· | |
| M | 5768 | hCoV-19/England/204221440/2020\|EPI_ISL_593813\|2020 | ···················· | ···················· | ····················· | ···················· | ···················· | ···················· | |
| M | 5769 | hCoV-19/England/204221441/2020\|EPI_ISL_593814\|2020 | ···················· | ···················· | ····················· | ···················· | ···················· | ···················· | |
| M | 5770 | hCoV-19/England/204221444/2020\|EPI_ISL_593815\|2020 | ···················· | ···················· | ····················· | ···················· | ···················· | ···················· | |
| M | 5771 | hCoV-19/England/204221445/2020\|EPI_ISL_593816\|2020 | ···················· | ···················· | ····················· | ···················· | ···················· | ···················· | |
| M | 5772 | hCoV-19/England/204221446/2020\|EPI_ISL_593817\|2020 | ···················· | ···················· | ····················· | ···················· | ···················· | ···················· | |
| M | 5773 | hCoV-19/England/204221448/2020\|EPI_ISL_593818\|2020 | ···················· | ···················· | ····················· | ···················· | ···················· | ···················· | |
| M | 5774 | hCoV-19/England/204221451/2020\|EPI_ISL_593819\|2020 | ···················· | ···················· | ····················· | ···················· | ···················· | ···················· | |
| M | 5775 | hCoV-19/England/204221453/2020\|EPI_ISL_593820\|2020 | ···················· | ···················· | ····················· | ···················· | ···················· | ···················· | |
| M | 5776 | hCoV-19/England/204221455/2020\|EPI_ISL_593822\|2020 | ···················· | ···················· | ····················· | ···················· | ···················· | ···················· | |
| M | 5777 | hCoV-19/England/204221459/2020\|EPI_ISL_593824\|2020 | ···················· | ···················· | ····················· | ···················· | ···················· | ···················· | |
| M | 5778 | hCoV-19/England/204221460/2020\|EPI_ISL_593825\|2020 | ···················· | ···················· | ····················· | ···················· | ···················· | ···················· | |
| M | 5779 | hCoV-19/England/204221461/2020\|EPI_ISL_593826\|2020 | ···················· | ···················· | ····················· | ···················· | ···················· | ···················· | |
| M | 5780 | hCoV-19/England/204221462/2020\|EPI_ISL_593827\|2020 | ···················· | ···················· | ····················· | ···················· | ···················· | ···················· | |
| M | 5781 | hCoV-19/England/204221463/2020\|EPI_ISL_593828\|2020 | ···················· | ···················· | ····················· | ···················· | ···················· | ···················· | |
| M | 5782 | hCoV-19/England/204221501/2020\|EPI_ISL_593829\|2020 | ···················· | ···················· | ····················· | ···················· | ···················· | ···················· | |
| M | 5783 | hCoV-19/England/204221504/2020\|EPI_ISL_593832\|2020 | ···················· | ···················· | ····················· | ···················· | ···················· | ···················· | |
| M | 5784 | hCoV-19/England/204221506/2020\|EPI_ISL_593833\|2020 | ···················· | ···················· | ····················· | ···················· | ···················· | ···················· | |
| M | 5785 | hCoV-19/England/204221510/2020\|EPI_ISL_593834\|2020 | ···················· | ···················· | ····················· | ···················· | ···················· | ···················· | |

| | | | | | | | |
|---|---|---|---|---|---|---|---|
| M | 5786 hCoV-19/England/204221511/2020\|EPI_ISL_593835\|2020 | ···················· | ···················· | ····················· | ···················· | ···················· | ···················· |
| M | 5787 hCoV-19/England/204221512/2020\|EPI_ISL_593836\|2020 | ···················· | ···················· | ····················· | ···················· | ···················· | ···················· |
| M | 5788 hCoV-19/England/204221516/2020\|EPI_ISL_593839\|2020 | ···················· | ···················· | ····················· | ···················· | ···················· | ···················· |
| M | 5789 hCoV-19/England/204221517/2020\|EPI_ISL_593840\|2020 | ···················· | ···················· | ····················· | ···················· | ···················· | ···················· |
| M | 5790 hCoV-19/England/204221518/2020\|EPI_ISL_593841\|2020 | ···················· | ···················· | ····················· | ···················· | ···················· | ···················· |
| M | 5791 hCoV-19/England/204221519/2020\|EPI_ISL_593842\|2020 | ···················· | ···················· | ····················· | ···················· | ···················· | ···················· |
| M | 5792 hCoV-19/England/204221520/2020\|EPI_ISL_593843\|2020 | ···················· | ···················· | ····················· | ···················· | ···················· | ···················· |
| M | 5793 hCoV-19/England/204221523/2020\|EPI_ISL_593844\|2020 | ···················· | ···················· | ····················· | ···················· | ···················· | ···················· |
| M | 5794 hCoV-19/England/204221524/2020\|EPI_ISL_593845\|2020 | ···················· | ···················· | ····················· | ···················· | ···················· | ···················· |
| M | 5795 hCoV-19/England/204221526/2020\|EPI_ISL_593847\|2020 | ···················· | ···················· | ····················· | ···················· | ···················· | ···················· |
| M | 5796 hCoV-19/England/204221527/2020\|EPI_ISL_593848\|2020 | ···················· | ···················· | ····················· | ···················· | ···················· | ···················· |
| M | 5797 hCoV-19/England/204221531/2020\|EPI_ISL_593851\|2020 | ···················· | ···················· | ····················· | ···················· | ···················· | ···················· |
| M | 5798 hCoV-19/France/OCC-40/2020\|EPI_ISL_593855\|2020-06- | ···················· | ···················· | ····················· | ···················· | ···················· | ···················· |
| M | 5799 hCoV-19/France/OCC-41/2020\|EPI_ISL_593856\|2020-06- | ···················· | ···················· | ····················· | ···················· | ···················· | ···················· |
| M | 5800 hCoV-19/France/OCC-42/2020\|EPI_ISL_593857\|2020-06- | ···················· | ···················· | ····················· | ···················· | ···················· | ···················· |
| M | 5801 hCoV-19/France/OCC-43/2020\|EPI_ISL_593858\|2020-06- | ···················· | ···················· | ····················· | ···················· | ···················· | ···················· |
| M | 5802 hCoV-19/France/OCC-44/2020\|EPI_ISL_593859\|2020-06- | ···················· | ···················· | ····················· | ···················· | ···················· | ···················· |
| M | 5803 hCoV-19/France/OCC-45/2020\|EPI_ISL_593860\|2020-06- | ···················· | ···················· | ····················· | ···················· | ···················· | ···················· |
| M | 5804 hCoV-19/France/OCC-46/2020\|EPI_ISL_593861\|2020-06- | ···················· | ···················· | ····················· | ···················· | ···················· | ···················· |
| M | 5805 hCoV-19/France/OCC-47/2020\|EPI_ISL_593862\|2020-06- | ···················· | ···················· | ····················· | ···················· | ···················· | ···················· |
| M | 5806 hCoV-19/France/OCC-48/2020\|EPI_ISL_593863\|2020-06- | ···················· | ···················· | ····················· | ···················· | ···················· | ···················· |
| M | 5807 hCoV-19/France/OCC-49/2020\|EPI_ISL_593864\|2020-06- | ···················· | ···················· | ····················· | ···················· | ···················· | ···················· |
| M | 5808 hCoV-19/France/OCC-50/2020\|EPI_ISL_593865\|2020-06- | ···················· | ···················· | ····················· | ···················· | ···················· | ···················· |
| M | 5809 hCoV-19/France/OCC-51/2020\|EPI_ISL_593866\|2020-06- | ···················· | ···················· | ····················· | ···················· | ···················· | ···················· |
| M | 5810 hCoV-19/France/OCC-52/2020\|EPI_ISL_593867\|2020-07- | ···················· | ···················· | ····················· | ···················· | ···················· | ···················· |
| M | 5811 hCoV-19/France/OCC-53/2020\|EPI_ISL_593868\|2020-07- | ···················· | ···················· | ····················· | ···················· | ···················· | ···················· |
| M | 5812 hCoV-19/France/OCC-54/2020\|EPI_ISL_593869\|2020-07- | ···················· | ···················· | ····················· | ···················· | ···················· | ···················· |
| M | 5813 hCoV-19/France/OCC-55/2020\|EPI_ISL_593870\|2020-07- | ···················· | ·A·················· | ····················· | ···················· | ···················· | ··A················· |
| M | 5814 hCoV-19/France/OCC-56/2020\|EPI_ISL_593871\|2020-07- | ···················· | ···················· | ····················· | ···················· | ···················· | ···················· |
| M | 5815 hCoV-19/France/OCC-57/2020\|EPI_ISL_593872\|2020-07- | ···················· | ···················· | ····················· | ···················· | ···················· | ···················· |
| M | 5816 hCoV-19/France/OCC-58/2020\|EPI_ISL_593873\|2020-07- | ···················· | ···················· | ····················· | ···················· | ···················· | ···················· |
| M | 5817 hCoV-19/France/OCC-59/2020\|EPI_ISL_593874\|2020-07- | ···················· | ···················· | ····················· | ···················· | ···················· | ···················· |
| M | 5818 hCoV-19/France/OCC-60/2020\|EPI_ISL_593875\|2020-07- | ···················· | ···················· | ····················· | ···················· | ···················· | ···················· |
| M | 5819 hCoV-19/France/OCC-62/2020\|EPI_ISL_593877\|2020-07- | ···················· | ···················· | ····················· | ···················· | ···················· | ···················· |
| M | 5820 hCoV-19/France/OCC-63/2020\|EPI_ISL_593878\|2020-08- | ···················· | ···················· | ····················· | ···················· | ···················· | ···················· |
| M | 5821 hCoV-19/France/OCC-64/2020\|EPI_ISL_593879\|2020-08- | ···················· | ···················· | ····················· | ···················· | ···················· | ···················· |
| M | 5822 hCoV-19/France/OCC-67/2020\|EPI_ISL_593881\|2020-06- | ···················· | ···················· | ····················· | ···················· | ···················· | ···················· |
| M | 5823 hCoV-19/France/OCC-68/2020\|EPI_ISL_593882\|2020-08- | ···················· | ···················· | ····················· | ···················· | ···················· | ···················· |
| M | 5824 hCoV-19/France/OCC-69/2020\|EPI_ISL_593883\|2020-07- | ···················· | ···················· | ····················· | ···················· | ···················· | ···················· |
| M | 5825 hCoV-19/France/OCC-70/2020\|EPI_ISL_593884\|2020-08- | ···················· | ···················· | ····················· | ···················· | ···················· | ···················· |
| M | 5826 hCoV-19/France/OCC-71/2020\|EPI_ISL_593885\|2020-08- | ···················· | ···················· | ····················· | ···················· | ···················· | ···················· |
| M | 5827 hCoV-19/France/OCC-72/2020\|EPI_ISL_593886\|2020-08- | ···················· | ···················· | ····················· | ···················· | ···················· | ···················· |
| M | 5828 hCoV-19/France/OCC-73/2020\|EPI_ISL_593887\|2020-08- | ···················· | ···A················ | ····················· | ···················· | ···················· | ···················· |
| M | 5829 hCoV-19/France/OCC-74/2020\|EPI_ISL_593888\|2020-09- | ···················· | ···A················ | ····················· | ···················· | ···················· | ···················· |
| M | 5830 hCoV-19/France/OCC-75/2020\|EPI_ISL_593889\|2020-08- | ···················· | ···A················ | ····················· | ···················· | ···················· | ···················· |
| M | 5831 hCoV-19/France/OCC-79/2020\|EPI_ISL_593890\|2020-06- | ···················· | ···················· | ····················· | ···················· | ···················· | ···················· |
| M | 5832 hCoV-19/France/OCC-80/2020\|EPI_ISL_593891\|2020-06- | ···················· | ···················· | ····················· | ···················· | ···················· | ···················· |
| M | 5833 hCoV-19/England/QEUH-A4CF4D/2020\|EPI_ISL_596070\|20 | ···················· | ···················· | ····················· | ···················· | ···················· | ···················· |
| M | 5834 hCoV-19/England/QEUH-A4CF6B/2020\|EPI_ISL_596071\|20 | ···················· | ···················· | ····················· | ···················· | ···················· | ···················· |
| M | 5835 hCoV-19/England/QEUH-A4CF7A/2020\|EPI_ISL_596072\|20 | ···················· | ···················· | ····················· | ···················· | ···················· | ···················· |
| M | 5836 hCoV-19/England/QEUH-A4CF98/2020\|EPI_ISL_596073\|20 | ···················· | ···················· | ····················· | ···················· | ···················· | ···················· |
| M | 5837 hCoV-19/England/QEUH-A4CFA7/2020\|EPI_ISL_596074\|20 | ···················· | ···················· | ····················· | ···················· | ···················· | ···················· |
| M | 5838 hCoV-19/England/QEUH-A4CFB6/2020\|EPI_ISL_596075\|20 | ···················· | ···················· | ····················· | ···················· | ···················· | ···················· |
| M | 5840 hCoV-19/England/QEUH-A4CFD4/2020\|EPI_ISL_596077\|20 | ···················· | ···················· | ····················· | ···················· | ···················· | ···················· |
| M | 5841 hCoV-19/England/QEUH-A4CFE3/2020\|EPI_ISL_596078\|20 | ···················· | ···················· | ····················· | ···················· | ···················· | ···················· |
| M | 5842 hCoV-19/England/QEUH-A4CFF2/2020\|EPI_ISL_596079\|20 | ···················· | ···················· | ····················· | ···················· | ···················· | ···················· |
| M | 5843 hCoV-19/England/QEUH-A4D00D/2020\|EPI_ISL_596080\|20 | ···················· | ···················· | ····················· | ···················· | ···················· | ···················· |
| M | 5844 hCoV-19/England/QEUH-A4D01C/2020\|EPI_ISL_596081\|20 | ···················· | ···················· | ····················· | ···················· | ···················· | ···················· |
| M | 5845 hCoV-19/England/QEUH-A4D02B/2020\|EPI_ISL_596082\|20 | ···················· | ···················· | ····················· | ···················· | ···················· | ···················· |
| M | 5846 hCoV-19/England/QEUH-A4D067/2020\|EPI_ISL_596083\|20 | ···················· | ···················· | ····················· | ···················· | ···················· | ···················· |
| M | 5847 hCoV-19/England/QEUH-A4D076/2020\|EPI_ISL_596084\|20 | ···················· | ·A·················· | ····················· | ···················· | ···················· | ··A················· |
| M | 5848 hCoV-19/England/QEUH-A4D085/2020\|EPI_ISL_596085\|20 | ···················· | ···················· | ····················· | ···················· | ···················· | ···················· |
| M | 5849 hCoV-19/England/QEUH-A4D0A3/2020\|EPI_ISL_596087\|20 | ···················· | ···················· | ····················· | ···················· | ···················· | ···················· |
| M | 5850 hCoV-19/England/QEUH-A4D0B2/2020\|EPI_ISL_596088\|20 | ···················· | ···················· | ····················· | ···················· | ···················· | ···················· |
| M | 5851 hCoV-19/England/QEUH-A4D0C1/2020\|EPI_ISL_596089\|20 | ···················· | ···················· | ····················· | ···················· | ···················· | ···················· |
| M | 5852 hCoV-19/England/QEUH-A4D0D0/2020\|EPI_ISL_596090\|20 | ···················· | ···················· | ····················· | ···················· | ···················· | ···················· |
| M | 5853 hCoV-19/England/QEUH-A4D10A/2020\|EPI_ISL_596091\|20 | ···················· | ···················· | ····················· | ···················· | ···················· | ···················· |
| M | 5854 hCoV-19/England/QEUH-A4D119/2020\|EPI_ISL_596092\|20 | ···················· | ···················· | ····················· | ···················· | ···················· | ···················· |
| M | 5855 hCoV-19/England/QEUH-A4D128/2020\|EPI_ISL_596093\|20 | ···················· | ···················· | ····················· | ···················· | ···················· | ···················· |
| M | 5856 hCoV-19/England/QEUH-A4D146/2020\|EPI_ISL_596095\|20 | ···················· | ···················· | ····················· | ···················· | ···················· | ···················· |
| M | 5857 hCoV-19/England/QEUH-A4D155/2020\|EPI_ISL_596096\|20 | ···················· | ···················· | ····················· | ···················· | ···················· | ···················· |
| M | 5858 hCoV-19/Scotland/QEUH-A4D1A0/2020\|EPI_ISL_596099\|2 | ···················· | ···················· | ····················· | ···················· | ···················· | ···················· |
| M | 5859 hCoV-19/England/QEUH-A4D234/2020\|EPI_ISL_596103\|20 | ···················· | ···················· | ····················· | ···················· | ···················· | ···················· |
| M | 5860 hCoV-19/Scotland/QEUH-A4D243/2020\|EPI_ISL_596104\|2 | ···················· | ···················· | ····················· | ···················· | ···················· | ···················· |
| M | 5861 hCoV-19/England/QEUH-A4D252/2020\|EPI_ISL_596105\|20 | ···················· | ···················· | ····················· | ···················· | ···················· | ···················· |
| M | 5862 hCoV-19/Scotland/QEUH-A4D29E/2020\|EPI_ISL_596106\|2 | ···················· | ···················· | ····················· | ···················· | ···················· | ···················· |
| M | 5863 hCoV-19/England/QEUH-A4D2CB/2020\|EPI_ISL_596107\|20 | ···················· | ···················· | ····················· | ···················· | ···················· | ···················· |
| M | 5864 hCoV-19/England/QEUH-A4D2F8/2020\|EPI_ISL_596108\|20 | ···················· | ···················· | ····················· | ···················· | ···················· | ···················· |
| M | 5865 hCoV-19/England/QEUH-A4D3B9/2020\|EPI_ISL_596110\|20 | ···················· | ···················· | ····················· | ···················· | ···················· | ···················· |
| M | 5866 hCoV-19/England/QEUH-A4D3C8/2020\|EPI_ISL_596111\|20 | ···················· | ···················· | ····················· | ···················· | ···················· | ···················· |

```
M    5867 hCoV-19/England/QEUH-A4D3D7/2020|EPI_ISL_596112|20    ....................  ....................  ....................  ....................  ....................  ....................
M    5868 hCoV-19/England/QEUH-A6361E/2020|EPI_ISL_596124|20    ....................  ....................  ....................  ....................  ....................  ....................
M    5869 hCoV-19/England/QEUH-A6362D/2020|EPI_ISL_596125|20    ....................  ....................  ....................  ....................  ....................  ....................
M    5870 hCoV-19/England/QEUH-A6364B/2020|EPI_ISL_596126|20    ....................  ....................  ....................  ....................  ....................  ....................
M    5871 hCoV-19/England/QEUH-A6365A/2020|EPI_ISL_596127|20    ....................  ....................  ....................  ....................  ....................  ....................
M    5872 hCoV-19/Scotland/QEUH-A63669/2020|EPI_ISL_596128|2    ....................  ....................  ....................  ....................  ....................  ....................
M    5873 hCoV-19/Scotland/QEUH-A63678/2020|EPI_ISL_596129|2    ....................  ....................  ....................  ....................  ....................  ....................
M    5874 hCoV-19/Scotland/QEUH-A63687/2020|EPI_ISL_596130|2    ....................  ....................  ....................  ....................  ....................  ....................
M    5875 hCoV-19/England/QEUH-A63696/2020|EPI_ISL_596131|20    ....................  ....................  ....................  ....................  ....................  ....................
M    5876 hCoV-19/England/QEUH-A636A5/2020|EPI_ISL_596132|20    ....................  ....................  ....................  ....................  ....................  ....................
M    5877 hCoV-19/Scotland/QEUH-A636B4/2020|EPI_ISL_596133|2    ....................  ....................  ....................  ....................  ....................  ....................
M    5878 hCoV-19/Scotland/QEUH-A636C3/2020|EPI_ISL_596134|2    ....................  ....................  ....................  ....................  ....................  ....................
M    5879 hCoV-19/England/QEUH-A636E1/2020|EPI_ISL_596135|20    ....................  ....................  ....................  ....................  ....................  ....................
M    5880 hCoV-19/Scotland/QEUH-A6370C/2020|EPI_ISL_596136|2    ....................  ....................  ....................  ....................  ....................  ....................
M    5881 hCoV-19/Scotland/QEUH-A6371B/2020|EPI_ISL_596137|2    ....................  ....................  ....................  ....................  ....................  ....................
M    5883 hCoV-19/Scotland/QEUH-A63748/2020|EPI_ISL_596140|2    ....................  ....................  ....................  ....................  ....................  ....................
M    5884 hCoV-19/Scotland/QEUH-A63757/2020|EPI_ISL_596141|2    ....................  ....................  ....................  ....................  ....................  ....................
M    5886 hCoV-19/Scotland/QEUH-A63775/2020|EPI_ISL_596143|2    ....................  ....................  ....................  ....................  ....................  ....................
M    5887 hCoV-19/Scotland/QEUH-A637A2/2020|EPI_ISL_596145|2    ....................  ....................  ....................  ....................  ....................  ....................
M    5888 hCoV-19/Scotland/QEUH-A637B1/2020|EPI_ISL_596146|2    ....................  ....................  ....................  ....................  ....................  ....................
M    5889 hCoV-19/England/ALDP-9E9295/2020|EPI_ISL_584196|20    ....................  ....................  ....................  ....................  ....................  ....................
M    5890 hCoV-19/England/ALDP-9E92A4/2020|EPI_ISL_584197|20    ....................  ....................  ....................  ....................  ....................  ....................
M    5891 hCoV-19/England/ALDP-9E92C2/2020|EPI_ISL_584199|20    ....................  ....................  ....................  ....................  ....................  ....................
M    5892 hCoV-19/Scotland/CVR4541/2020|EPI_ISL_585400|2020-    ....................  ....................  ....................  ....................  ....................  ....................
M    5894 hCoV-19/Scotland/CVR4544/2020|EPI_ISL_585403|2020-    ....................  ....................  ....................  ....................  ....................  ....................
M    5895 hCoV-19/Scotland/CVR4545/2020|EPI_ISL_585404|2020-    ....................  ....................  ....................  ....................  ....................  ....................
M    5896 hCoV-19/Scotland/CVR4546/2020|EPI_ISL_585405|2020-    ....................  ....................  ....................  ....................  ....................  ....................
M    5897 hCoV-19/Scotland/CVR4547/2020|EPI_ISL_585406|2020-    ....................  ....................  ....................  ....................  ....................  ....................
M    5898 hCoV-19/Scotland/CVR4548/2020|EPI_ISL_585407|2020-    ....................  ....................  ....................  ....................  ....................  ....................
M    5899 hCoV-19/Scotland/CVR4551/2020|EPI_ISL_585410|2020-    ....................  ....................  ....................  ....................  ....................  ....................
M    5900 hCoV-19/Scotland/CVR4552/2020|EPI_ISL_585411|2020-    ....................  ....................  ....................  ....................  ....................  ....................
M    5901 hCoV-19/Scotland/CVR4553/2020|EPI_ISL_585412|2020-    ....................  ....................  ....................  ....................  ....................  ....................
M    5902 hCoV-19/Scotland/CVR4555/2020|EPI_ISL_585413|2020-    ....................  ....................  ....................  ....................  ....................  ....................
M    5903 hCoV-19/Scotland/CVR4556/2020|EPI_ISL_585414|2020-    ....................  ....................  ....................  ....................  ....................  ....................
M    5904 hCoV-19/Scotland/CVR4559/2020|EPI_ISL_585417|2020-    ....................  ....................  ....................  ....................  ....................  ....................
M    5905 hCoV-19/Scotland/CVR4560/2020|EPI_ISL_585418|2020-    ....................  ....................  ....................  ....................  ....................  ....................
M    5906 hCoV-19/Scotland/CVR4561/2020|EPI_ISL_585419|2020-    ....................  ....................  ....................  ....................  ....................  ....................
M    5907 hCoV-19/Scotland/CVR4562/2020|EPI_ISL_585420|2020-    ....................  ....................  ....................  ....................  ....................  ....................
M    5908 hCoV-19/Scotland/CVR4621/2020|EPI_ISL_585422|2020-    ....................  ....................  ....................  ....................  ....................  ....................
M    5909 hCoV-19/Scotland/CVR4623/2020|EPI_ISL_585423|2020-    ....................  ....................  ....................  ....................  ....................  ....................
M    5910 hCoV-19/Scotland/CVR4732/2020|EPI_ISL_585425|2020-    ....................  ....................  ....................  ....................  ....................  ....................
M    5911 hCoV-19/Scotland/EDB7866/2020|EPI_ISL_585426|2020-    ....................  ....................  ....................  ....................  ....................  ....................
M    5912 hCoV-19/Scotland/EDB7871/2020|EPI_ISL_585427|2020-    ....................  ....................  ....................  ....................  ....................  ....................
M    5913 hCoV-19/Scotland/EDB7874/2020|EPI_ISL_585428|2020-    ....................  ....................  ....................  ....................  ....................  ....................
M    5914 hCoV-19/Scotland/EDB7876/2020|EPI_ISL_585429|2020-    ....................  ....................  ....................  ....................  ....................  ....................
M    5915 hCoV-19/Scotland/EDB7877/2020|EPI_ISL_585430|2020-    ....................  ....................  ....................  ....................  ....................  ....................
M    5916 hCoV-19/Scotland/EDB7878/2020|EPI_ISL_585431|2020-    ....................  ....................  ....................  ....................  ....................  ....................
M    5917 hCoV-19/Scotland/EDB7879/2020|EPI_ISL_585432|2020-    ....................  ....................  ....................  ....................  ....................  ....................
M    5918 hCoV-19/Scotland/EDB7880/2020|EPI_ISL_585433|2020-    ....................  ....................  ....................  ....................  ....................  ....................
M    5919 hCoV-19/Scotland/EDB7881/2020|EPI_ISL_585434|2020-    ....................  ....................  ....................  ....................  ....................  ....................
M    5920 hCoV-19/Scotland/EDB7882/2020|EPI_ISL_585435|2020-    ....................  ....................  ....................  ....................  ....................  ....................
M    5921 hCoV-19/Scotland/EDB7883/2020|EPI_ISL_585436|2020-    ....................  ....................  ....................  ....................  ....................  ....................
M    5922 hCoV-19/Scotland/EDB7885/2020|EPI_ISL_585438|2020-    ....................  ....................  ....................  ....................  ....................  ....................
M    5923 hCoV-19/Scotland/EDB7886/2020|EPI_ISL_585439|2020-    ....................  ....................  ....................  ....................  ....................  ....................
M    5924 hCoV-19/Scotland/EDB7887/2020|EPI_ISL_585440|2020-    ....................  ....................  ....................  ....................  ....................  ....................
M    5925 hCoV-19/Scotland/EDB7891/2020|EPI_ISL_585441|2020-    ....................  ....................  ....................  ....................  ....................  ....................
M    5926 hCoV-19/Scotland/EDB7895/2020|EPI_ISL_585443|2020-    ....................  ....................  ....................  ....................  ....................  ....................
M    5927 hCoV-19/Scotland/EDB7909/2020|EPI_ISL_585445|2020-    ....................  ....................  ....................  ....................  ....................  ....................
M    5928 hCoV-19/Scotland/EDB7910/2020|EPI_ISL_585446|2020-    ....................  ....................  ....................  ....................  ....................  ....................
M    5929 hCoV-19/Scotland/EDB7915/2020|EPI_ISL_585448|2020-    ....................  ....................  ....................  ....................  ....................  ....................
M    5930 hCoV-19/Scotland/EDB7916/2020|EPI_ISL_585449|2020-    ....................  ....................  ....................  ....................  ....................  ....................
M    5931 hCoV-19/Scotland/EDB7917/2020|EPI_ISL_585450|2020-    ....................  ....................  ....................  ....................  ......T.............  ....................
M    5932 hCoV-19/Scotland/EDB7924/2020|EPI_ISL_585452|2020-    ....................  ....................  ....................  ....................  ....................  ....................
M    5933 hCoV-19/Scotland/EDB7926/2020|EPI_ISL_585453|2020-    ....................  ....................  ....................  ....................  ....................  ....................
M    5934 hCoV-19/Scotland/EDB7927/2020|EPI_ISL_585454|2020-    ....................  ....................  ....................  ....................  ....................  ....................
M    5935 hCoV-19/Scotland/EDB7986/2020|EPI_ISL_585466|2020-    ....................  ....................  ....................  ....................  ....................  ....................
M    5938 hCoV-19/Scotland/EDB8031/2020|EPI_ISL_585472|2020-    ....................  ....................  ....................  ....................  ....................  ....................
M    5939 hCoV-19/Scotland/EDB8032/2020|EPI_ISL_585473|2020-    ....................  ....................  ....................  ....................  ....................  ....................
M    5940 hCoV-19/Scotland/EDB8090/2020|EPI_ISL_585476|2020-    ....................  ....................  ....................  ....................  ....................  ....................
M    5941 hCoV-19/Scotland/EDB8249/2020|EPI_ISL_585499|2020-    ....................  ....................  ....................  ....................  ....................  ....................
M    5942 hCoV-19/England/ALDP-A2E03F/2020|EPI_ISL_586700|20    ....................  ....................  ....................  ....................  ....................  ....................
M    5943 hCoV-19/England/ALDP-A2D9F9/2020|EPI_ISL_586701|20    ....................  ....................  ....................  ....................  ....................  ....................
M    5944 hCoV-19/England/ALDP-A2E309/2020|EPI_ISL_586702|20    ....................  ....................  ....................  ....................  ....................  ....................
M    5945 hCoV-19/England/ALDP-A2E1D2/2020|EPI_ISL_586703|20    ....................  ....................  ....................  ....................  ....................  ....................
M    5946 hCoV-19/England/ALDP-A2E363/2020|EPI_ISL_586704|20    ....................  ....................  ....................  ....................  ......T.............  ....................
M    5947 hCoV-19/England/ALDP-A2D3EC/2020|EPI_ISL_586705|20    ....................  ....................  ....................  ....................  ....................  ....................
M    5948 hCoV-19/England/ALDP-A2DFE8/2020|EPI_ISL_586706|20    ....................  ....................  ....................  ....................  ....................  ....................
M    5949 hCoV-19/England/ALDP-A2D3CE/2020|EPI_ISL_586707|20    ....................  ....................  ....................  ....................  ....................  ....................
M    5950 hCoV-19/England/ALDP-A2E381/2020|EPI_ISL_586710|20    ....................  ....................  ....................  ....................  ....................  ....................
M    5951 hCoV-19/England/ALDP-A2E3DC/2020|EPI_ISL_586711|20    ....................  ....................  ....................  ....................  ....................  ....................
```

M 5952 hCoV-19/England/ALDP-A2D328/2020|EPI_ISL_586712|20 ···················· ···················· ····················· ···················· ···················· ····················
M 5953 hCoV-19/England/ALDP-A2D4AD/2020|EPI_ISL_586713|20 ···················· ···················· ····················· ···················· ···················· ····················
M 5954 hCoV-19/England/ALDP-A2D07C/2020|EPI_ISL_586714|20 ···················· ···················· ····················· ···················· ···················· ····················
M 5955 hCoV-19/England/ALDP-A2D0E5/2020|EPI_ISL_586715|20 ···················· ···················· ····················· ···················· ···················· ····················
M 5956 hCoV-19/England/ALDP-A2D917/2020|EPI_ISL_586716|20 ···················· ···················· ····················· ···················· ···················· ····················
M 5957 hCoV-19/England/ALDP-A2E3FA/2020|EPI_ISL_586717|20 ···················· ·A·················· ····················· ···················· ···················· ····················
M 5958 hCoV-19/England/ALDP-A2E47F/2020|EPI_ISL_586718|20 ···················· ···················· ····················· ···················· ···················· ····················
M 5959 hCoV-19/England/ALDP-A2E002/2020|EPI_ISL_586719|20 ···················· ···················· ····················· ···················· ···················· ····················
M 5960 hCoV-19/England/ALDP-A2E80A/2020|EPI_ISL_586720|20 ···················· ···················· ····················· ···················· ···················· ····················
M 5961 hCoV-19/England/ALDP-A2DDDF/2020|EPI_ISL_586721|20 ···················· ···················· ····················· ···················· ···················· ····················
M 5962 hCoV-19/England/ALDP-A2DBA8/2020|EPI_ISL_586722|20 ···················· ···················· ····················· ···················· ···················· ····················
M 5963 hCoV-19/England/ALDP-A2E697/2020|EPI_ISL_586723|20 ···················· ···················· ····················· ···················· ···················· ····················
M 5964 hCoV-19/England/ALDP-A2DB3F/2020|EPI_ISL_586724|20 ···················· ···················· ····················· ···················· ···················· ····················
M 5965 hCoV-19/England/ALDP-A2D874/2020|EPI_ISL_586726|20 ···················· ···················· ····················· ···················· ···················· ····················
M 5966 hCoV-19/England/ALDP-A2DF33/2020|EPI_ISL_586727|20 ···················· ···················· ····················· ···················· ···················· ····················
M 5967 hCoV-19/England/ALDP-A2DF24/2020|EPI_ISL_586729|20 ···················· ···················· ····················· ···················· ···················· ····················
M 5968 hCoV-19/England/ALDP-A2DC5A/2020|EPI_ISL_586730|20 ···················· ···················· ····················· ···················· ···················· ····················
M 5969 hCoV-19/England/ALDP-A2D698/2020|EPI_ISL_586731|20 ···················· ···················· ····················· ···················· ···················· ····················
M 5970 hCoV-19/England/ALDP-A2DE36/2020|EPI_ISL_586732|20 ···················· ···················· ····················· ···················· ···················· ····················
M 5971 hCoV-19/England/ALDP-A2D9AE/2020|EPI_ISL_586733|20 ···················· ···················· ····················· ···················· ···················· ····················
M 5972 hCoV-19/England/ALDP-A2D62F/2020|EPI_ISL_586734|20 ···················· ·············C······ ····················· ···················· ···················· ····················
M 5973 hCoV-19/England/ALDP-A2DEBE/2020|EPI_ISL_586735|20 ···················· ···················· ····················· ···················· ···················· ····················
M 5974 hCoV-19/England/ALDP-A2D962/2020|EPI_ISL_586736|20 ···················· ···················· ····················· ···················· ···················· ····················
M 5975 hCoV-19/England/ALDP-A2D22B/2020|EPI_ISL_586737|20 ···················· ···················· ····················· ···················· ···················· ····················
M 5976 hCoV-19/England/ALDP-A2E336/2020|EPI_ISL_586738|20 ···················· ·A·················· ····················· ···················· ···················· ····················
M 5977 hCoV-19/England/ALDP-A2DF8E/2020|EPI_ISL_586739|20 ···················· ···················· ····················· ···················· ···················· ····················
M 5978 hCoV-19/England/ALDP-A2E530/2020|EPI_ISL_586740|20 ···················· ···················· ····················· ···················· ···················· ····················
M 5979 hCoV-19/England/ALDP-A2E7FE/2020|EPI_ISL_586741|20 ···················· ···················· ····················· ···················· ···················· ····················
M 5980 hCoV-19/England/ALDP-A2E776/2020|EPI_ISL_586742|20 ···················· ···················· ····················· ···················· ···················· ····················
M 5981 hCoV-19/England/ALDP-A2E275/2020|EPI_ISL_586743|20 ···················· ···················· ····················· ···················· ···T················ ····················
M 5982 hCoV-19/England/ALDP-A2D6B6/2020|EPI_ISL_586744|20 ···················· ···················· ····················· ···················· ···················· ····················
M 5983 hCoV-19/England/ALDP-A2D407/2020|EPI_ISL_586745|20 ···················· ···················· ····················· ···················· ···················· ····················
M 5984 hCoV-19/England/ALDP-A2E12D/2020|EPI_ISL_586746|20 ···················· ···················· ····················· ···················· ···················· ····················
M 5985 hCoV-19/England/ALDP-A2DC78/2020|EPI_ISL_586747|20 ···················· ···················· ····················· ···················· ···················· ····················
M 5986 hCoV-19/England/ALDP-A2DD0C/2020|EPI_ISL_586748|20 ···················· ···················· ····················· ···················· ···················· ····················
M 5987 hCoV-19/England/ALDP-A2DC3C/2020|EPI_ISL_586749|20 ···················· ···················· ····················· ···················· ···················· ····················
M 5988 hCoV-19/England/ALDP-A2D1D3/2020|EPI_ISL_586750|20 ···················· ···················· ····················· ···················· ···················· ····················
M 5989 hCoV-19/England/ALDP-A2DB20/2020|EPI_ISL_586751|20 ···················· ···················· ····················· ···················· ···················· ····················
M 5990 hCoV-19/England/ALDP-A2E521/2020|EPI_ISL_586752|20 ···················· ···················· ····················· ···················· ···················· ····················
M 5991 hCoV-19/England/ALDP-A2E390/2020|EPI_ISL_586755|20 ···················· ···················· ····················· ···················· ···················· ····················
M 5992 hCoV-19/England/ALDP-A2D856/2020|EPI_ISL_586756|20 ···················· ·A·················· ····················· ···················· ···················· ··A·················
M 5993 hCoV-19/England/ALDP-A2E58B/2020|EPI_ISL_586757|20 ···················· ···················· ····················· ···················· ···················· ····················
M 5994 hCoV-19/England/ALDP-A2D953/2020|EPI_ISL_586758|20 ···················· ···················· ····················· ···················· ···················· ····················
M 5995 hCoV-19/England/ALDP-A2DA7E/2020|EPI_ISL_586759|20 ···················· ···················· ····················· ···················· ···················· ····················
M 5996 hCoV-19/England/ALDP-A2D7C2/2020|EPI_ISL_586760|20 ···················· ···················· ····················· ···················· ···················· ····················
M 5997 hCoV-19/England/ALDP-A2D06D/2020|EPI_ISL_586761|20 ···················· ···················· ····················· ···················· ···················· ····················
M 5998 hCoV-19/England/ALDP-A2D2A3/2020|EPI_ISL_586762|20 ···················· ···················· ····················· ···················· ···················· ····················
M 5999 hCoV-19/England/ALDP-A2D3A0/2020|EPI_ISL_586763|20 ···················· ···················· ····················· ···················· ···················· ····················
M 6000 hCoV-19/England/ALDP-A2D9EA/2020|EPI_ISL_586764|20 ···················· ···················· ····················· ···················· ···················· ····················
M 6001 hCoV-19/England/ALDP-A2D56E/2020|EPI_ISL_586765|20 ···················· ···················· ····················· ···················· ···················· ····················
M 6002 hCoV-19/England/ALDP-A2E2FD/2020|EPI_ISL_586766|20 ···················· ···················· ····················· ···················· ···················· ····················
M 6003 hCoV-19/England/ALDP-A2DE63/2020|EPI_ISL_586767|20 ···················· ···················· ····················· ···················· ···················· ····················
M 6004 hCoV-19/England/ALDP-A2DA8D/2020|EPI_ISL_586768|20 ···················· ···················· ····················· ···················· ···················· ····················
M 6005 hCoV-19/England/ALDP-A2D355/2020|EPI_ISL_586769|20 ···················· ···················· ····················· ···················· ···················· ····················
M 6006 hCoV-19/England/ALDP-A2DD39/2020|EPI_ISL_586770|20 ···················· ···················· ····················· ···················· ···················· ····················
M 6007 hCoV-19/England/ALDP-A2D4BC/2020|EPI_ISL_586771|20 ···················· ···················· ····················· ···················· ···················· ····················
M 6008 hCoV-19/England/ALDP-A2D99F/2020|EPI_ISL_586773|20 ···················· ···················· ····················· ···················· ···················· ····················
M 6009 hCoV-19/England/ALDP-A2E5B8/2020|EPI_ISL_586774|20 ···················· ···················· ····················· ···················· ···················· ····················
M 6010 hCoV-19/England/ALDP-A2DA6F/2020|EPI_ISL_586775|20 ···················· ···················· ····················· ···················· ···················· ····················
M 6011 hCoV-19/England/ALDP-A2E819/2020|EPI_ISL_586776|20 ···················· ···················· ····················· ···················· ···················· ····················
M 6012 hCoV-19/England/ALDP-A2E6F1/2020|EPI_ISL_586777|20 ···················· ···················· ····················· ···················· ···················· ····················
M 6013 hCoV-19/England/ALDP-A2DF9D/2020|EPI_ISL_586778|20 ···················· ·A·················· ····················· ···················· ···················· ··A·················
M 6014 hCoV-19/England/ALDP-A2E679/2020|EPI_ISL_586779|20 ···················· ···················· ····················· ···················· ···················· ····················
M 6015 hCoV-19/England/ALDP-A2D12E/2020|EPI_ISL_586780|20 ···················· ···················· ····················· ···················· ···················· ····················
M 6016 hCoV-19/England/ALDP-A2E266/2020|EPI_ISL_586781|20 ···················· ···················· ····················· ···················· ···················· ····················
M 6017 hCoV-19/England/ALDP-A2E7A3/2020|EPI_ISL_586782|20 ···················· ·A·················· ····················· ···················· ···················· ··A·················
M 6018 hCoV-19/England/ALDP-A2D6F2/2020|EPI_ISL_586783|20 ···················· ···················· ····················· ···················· ···················· ····················
M 6019 hCoV-19/England/ALDP-A2D689/2020|EPI_ISL_586784|20 ···················· ···················· ····················· ···················· ···················· ····················
M 6020 hCoV-19/England/ALDP-A2DC1E/2020|EPI_ISL_586785|20 ···················· ···················· ····················· ···················· ···················· ····················
M 6021 hCoV-19/England/ALDP-A2DB5D/2020|EPI_ISL_586797|20 ···················· ···················· ····················· ···················· ···················· ····················
M 6022 hCoV-19/England/ALDP-A2DBE4/2020|EPI_ISL_586799|20 ···················· ···················· ····················· ···················· ···················· ····················
M 6023 hCoV-19/Canada/ON-S1465/2020|EPI_ISL_591103|2020-0 ···················· ···················· ····················· ···················· ···················· ····················
M 6024 hCoV-19/Canada/ON-S1474/2020|EPI_ISL_591110|2020-0 ···················· ···················· ····················· ···················· ···················· ····················
M 6025 hCoV-19/Canada/ON-S1476/2020|EPI_ISL_591112|2020-0 ···················· ···················· ····················· ···················· ···················· ····················
M 6026 hCoV-19/Canada/ON-S1481/2020|EPI_ISL_591115|2020-0 ···················· ···················· ····················· ···················· ···················· ····················
M 6027 hCoV-19/Canada/ON-S1482/2020|EPI_ISL_591116|2020-0 ···················· ···················· ····················· ···················· ···················· ····················
M 6028 hCoV-19/Canada/ON-S1483/2020|EPI_ISL_591117|2020-0 ···················· ···················· ····················· ···················· ···················· ····················
M 6029 hCoV-19/Canada/ON-S192/2020|EPI_ISL_591122|2020-03 ···················· ···················· ····················· ···················· ···················· ····················
M 6030 hCoV-19/USA/UT-UPHL-04306_2/2020|EPI_ISL_591297|20 ···················· ···················· ····················· ···················· ···················· ····················
M 6031 hCoV-19/USA/UT-UPHL-04307_2/2020|EPI_ISL_591298|20 ···················· ···················· ····················· ···················· ···················· ····················

M 6032 hCoV-19/Australia/VIC14255/2020|EPI_ISL_592420|202 ···················· ···················· ···················· ···················· ···················· ····················
M 6033 hCoV-19/Australia/VIC14257/2020|EPI_ISL_592421|202 ···················· ···················· ···················· ···················· ···················· ····················
M 6034 hCoV-19/Australia/VIC14258/2020|EPI_ISL_592422|202 ···················· ···················· ···················· ···················· ···················· ····················
M 6035 hCoV-19/Australia/VIC14262/2020|EPI_ISL_592423|202 ···················· ···················· ···················· ···················· ···················· ····················
M 6037 hCoV-19/Australia/VIC14264/2020|EPI_ISL_592425|202 ···················· ···················· ···················· ···················· ···················· ····················
M 6038 hCoV-19/Australia/VIC14265/2020|EPI_ISL_592426|202 ···················· ···················· ···················· ···················· ···················· ····················
M 6039 hCoV-19/Australia/VIC14266/2020|EPI_ISL_592427|202 ···················· ···················· ···················· ···················· ···················· ····················
M 6040 hCoV-19/Australia/VIC14272/2020|EPI_ISL_592428|202 ···················· ···················· ···················· ···················· ···················· ····················
M 6041 hCoV-19/Australia/VIC14276/2020|EPI_ISL_592429|202 ···················· ···················· ···················· ···················· ···················· ····················
M 6042 hCoV-19/Australia/VIC14285/2020|EPI_ISL_592435|202 ···················· ···················· ···················· ···················· ···················· ····················
M 6043 hCoV-19/Australia/VIC14296/2020|EPI_ISL_592440|202 ···················· ···················· ···················· ···················· ···················· ····················
M 6044 hCoV-19/Australia/VIC14298/2020|EPI_ISL_592441|202 ···················· ···················· ···················· ···················· ···················· ····················
M 6045 hCoV-19/Australia/VIC14299/2020|EPI_ISL_592442|202 ···················· ···················· ···················· ···················· ···················· ····················
M 6046 hCoV-19/Australia/VIC14300/2020|EPI_ISL_592443|202 ···················· ···················· ···················· ···················· ···················· ····················
M 6047 hCoV-19/Australia/VIC14306/2020|EPI_ISL_592444|202 ···················· ···················· ···················· ···················· ···················· ····················
M 6048 hCoV-19/Australia/VIC14308/2020|EPI_ISL_592445|202 ···················· ···················· ···················· ···················· ···················· ····················
M 6049 hCoV-19/Australia/VIC14312/2020|EPI_ISL_592446|202 ···················· ···················· ···················· ···················· ···················· ····················
M 6050 hCoV-19/Australia/VIC14315/2020|EPI_ISL_592447|202 ···················· ···················· ···················· ···················· ···················· ····················
M 6051 hCoV-19/Australia/VIC14316/2020|EPI_ISL_592448|202 ···················· ···················· ···················· ···················· ····S··············· ····················
M 6052 hCoV-19/Australia/VIC14320/2020|EPI_ISL_592449|202 ···················· ···················· ···················· ···················· ···················· ····················
M 6053 hCoV-19/Australia/VIC14323/2020|EPI_ISL_592450|202 ···················· ···················· ···················· ···················· ···················· ····················
M 6054 hCoV-19/Australia/VIC14327/2020|EPI_ISL_592451|202 ···················· ···················· ···················· ···················· ···················· ····················
M 6055 hCoV-19/Australia/VIC14329/2020|EPI_ISL_592452|202 ···················· ···················· ···················· ···················· ···················· ····················
M 6056 hCoV-19/Australia/VIC14331/2020|EPI_ISL_592453|202 ···················· ···················· ···················· ···················· ···················· ····················
M 6057 hCoV-19/Australia/VIC14332/2020|EPI_ISL_592454|202 ···················· ···················· ···················· ···················· ·········N·········· ····················
M 6058 hCoV-19/Australia/VIC14335/2020|EPI_ISL_592455|202 ···················· ···················· ···················· ···················· ···················· ····················
M 6059 hCoV-19/Australia/VIC14336/2020|EPI_ISL_592456|202 ···················· ···················· ···················· ···················· ···················· ····················
M 6061 hCoV-19/Australia/VIC14343/2020|EPI_ISL_592458|202 ···················· ···················· ···················· ···················· ···················· ····················
M 6062 hCoV-19/Australia/VIC14344/2020|EPI_ISL_592459|202 ···················· ···················· ···················· ···················· ···················· ····················
M 6063 hCoV-19/Australia/VIC14348/2020|EPI_ISL_592460|202 ···················· ···················· ···················· ···················· ···················· ····················
M 6065 hCoV-19/Australia/NSW1140/2020|EPI_ISL_593750|2020 ···················· ···················· ···················· ···················· ···················· ····················
M 6066 hCoV-19/Australia/NSW1141/2020|EPI_ISL_593751|2020 ···················· ···················· ···················· ···················· ···················· ····················
M 6067 hCoV-19/Australia/NSW1142/2020|EPI_ISL_593752|2020 ···················· ···················· ···················· ···················· ···················· ····················
M 6068 hCoV-19/Australia/NSW1143/2020|EPI_ISL_593753|2020 ···················· ·A·················· ···················· ···················· ···················· ···A················
M 6069 hCoV-19/Australia/NSW1145/2020|EPI_ISL_593754|2020 ···················· ···················· ···················· ···················· ···················· ····················
M 6070 hCoV-19/Australia/NSW1024/2020|EPI_ISL_593755|2020 ···················· ···················· ···················· ···················· ···················· ····················
M 6071 hCoV-19/Australia/NSW1027/2020|EPI_ISL_593756|2020 ···················· ···················· ···················· ···················· ···················· ····················
M 6072 hCoV-19/Australia/NSW1028/2020|EPI_ISL_593757|2020 ···················· ···················· ···················· ···················· ···················· ····················
M 6073 hCoV-19/Australia/NSW1029/2020|EPI_ISL_593758|2020 ···················· ···················· ···················· ···················· ···················· ····················
M 6074 hCoV-19/Australia/NSW1030/2020|EPI_ISL_593759|2020 ···················· ···················· ···················· ···················· ···················· ····················
M 6075 hCoV-19/Australia/NSW1086/2020|EPI_ISL_593766|2020 ···················· ···················· ···················· ···················· ···················· ····················
M 6076 hCoV-19/Australia/NSW1078/2020|EPI_ISL_593767|2020 ···················· ···················· ···················· ···················· ···················· ····················
M 6077 hCoV-19/Australia/NSW1114/2020|EPI_ISL_593770|2020 ········T··········· ···················· ···················· ···················· ···················· ····················
M 6078 hCoV-19/Australia/NSW1134/2020|EPI_ISL_593771|2020 ···················· ···················· ···················· ···················· ···················· ····················
M 6079 hCoV-19/Peru/LAM-HRL-239/2020|EPI_ISL_593773|2020- ···················· ···················· ···················· ···················· ···················· ····················
M 6080 hCoV-19/Peru/LAM-HRL-187/2020|EPI_ISL_593774|2020- ···················· ···················· ···················· ···················· ···················· ····················
M 6081 hCoV-19/Peru/LAM-HRL-223/2020|EPI_ISL_593775|2020- ···················· ···················· ···················· ···················· ···················· ····················
M 6082 hCoV-19/England/204221380/2020|EPI_ISL_593787|2020 ···················· ···················· ···················· ···················· ···················· ····················
M 6083 hCoV-19/England/204221390/2020|EPI_ISL_593790|2020 ···················· ···················· ···················· ···················· ···················· ····················
M 6084 hCoV-19/England/204221392/2020|EPI_ISL_593791|2020 ···················· ···················· ···················· ···················· ···················· ····················
M 6085 hCoV-19/England/204221397/2020|EPI_ISL_593792|2020 ···················· ···················· ···················· ···················· ···················· ····················
M 6086 hCoV-19/France/OCC-92/2020|EPI_ISL_593900|2020-10- ···················· ···················· ···················· ···················· ···················· ····················
M 6087 hCoV-19/France/OCC-93/2020|EPI_ISL_593901|2020-09- ···················· ···················· ···················· ···················· ···················· ····················
M 6088 hCoV-19/France/IDF-9113/2020|EPI_ISL_593902|2020-0 ···················· ···················· ···················· ···················· ···················· ····················
M 6089 hCoV-19/France/IDF-9132/2020|EPI_ISL_593903|2020-0 ···················· ···················· ···················· ···················· ···················· ····················
M 6090 hCoV-19/France/IDF-9135/2020|EPI_ISL_593904|2020-0 ···················· ···················· ···················· ···················· ···················· ····················
M 6091 hCoV-19/France/IDF-9137/2020|EPI_ISL_593905|2020-0 ···················· ···················· ···················· ···················· ···················· ····················
M 6092 hCoV-19/France/IDF-9139/2020|EPI_ISL_593906|2020-0 ···················· ···················· ···················· ···················· ···················· ····················
M 6093 hCoV-19/France/IDF-9141/2020|EPI_ISL_593907|2020-0 ···················· ···················· ···················· ···················· ···················· ····················
M 6094 hCoV-19/France/IDF-9142/2020|EPI_ISL_593908|2020-0 ···················· ···················· ···················· ···················· ···················· ····················
M 6095 hCoV-19/France/IDF-9143/2020|EPI_ISL_593909|2020-0 ···················· ···················· ···················· ···················· ···················· ····················
M 6096 hCoV-19/Scotland/QEUH-A4D1CE/2020|EPI_ISL_596100|2 ···················· ···················· ···················· ···················· ···················· ····················
M 6097 hCoV-19/Scotland/QEUH-A4D1DD/2020|EPI_ISL_596101|2 ···················· ···················· ···················· ···················· ···················· ····················
M 6098 hCoV-19/Scotland/QEUH-A4D225/2020|EPI_ISL_596102|2 ···················· ···················· ···················· ···················· ···················· ····················
M 6099 hCoV-19/England/QEUH-A4D3F5/2020|EPI_ISL_596114|20 ···················· ···················· ···················· ···················· ···················· ····················
M 6100 hCoV-19/England/QEUH-A4D401/2020|EPI_ISL_596115|20 ···················· ···················· ···················· ···················· ···················· ····················
M 6101 hCoV-19/England/QEUH-A63371/2020|EPI_ISL_596116|20 ···················· ···················· ···················· ···················· ···················· ····················
M 6102 hCoV-19/England/QEUH-A63380/2020|EPI_ISL_596117|20 ···················· ···················· ···················· ···················· ···················· ····················
M 6103 hCoV-19/England/QEUH-A633AE/2020|EPI_ISL_596118|20 ···················· ···················· ···················· ···················· ···················· ····················
M 6104 hCoV-19/England/QEUH-A633EA/2020|EPI_ISL_596119|20 ···················· ···················· ···················· ···················· ···················· ····················
M 6105 hCoV-19/England/QEUH-A6357B/2020|EPI_ISL_596120|20 ···················· ···················· ···················· ···················· ···················· ····················
M 6106 hCoV-19/England/QEUH-A635B7/2020|EPI_ISL_596121|20 ···················· ···················· ···················· ···················· ···················· ····················
M 6107 hCoV-19/England/QEUH-A635E4/2020|EPI_ISL_596122|20 ···················· ···················· ···················· ···················· ···················· ····················
M 6108 hCoV-19/England/QEUH-A635F3/2020|EPI_ISL_596123|20 ···················· ···················· ···················· ···················· ···················· ····················
M 6109 hCoV-19/Scotland/QEUH-A637C0/2020|EPI_ISL_596147|2 ···················· ···················· ···················· ···················· ···················· ····················
M 6110 hCoV-19/Scotland/QEUH-A637EE/2020|EPI_ISL_596149|2 ···················· ···················· ···················· ···················· ···················· ····················
M 6111 hCoV-19/Scotland/QEUH-A637FD/2020|EPI_ISL_596150|2 ···················· ···················· ···················· ···················· ···················· ····················
M 6112 hCoV-19/Scotland/QEUH-A63818/2020|EPI_ISL_596152|2 ···················· ···················· ···················· ···················· ···················· ····················
M 6113 hCoV-19/Scotland/QEUH-A63836/2020|EPI_ISL_596153|2 ···················· ···················· ···················· ···················· ···················· ····················
M 6114 hCoV-19/Scotland/QEUH-A63845/2020|EPI_ISL_596154|2 ···················· ···················· ···················· ···················· ···················· ····················

M 6115 hCoV-19/Scotland/QEUH-A63854/2020|EPI_ISL_596155|2 ···················· ···················· ···················· ···················· ···················· ····················
M 6116 hCoV-19/Scotland/QEUH-A63881/2020|EPI_ISL_596156|2 ···················· ···················· ···················· ···················· NNN················· ···················· 1
M 6118 hCoV-19/Scotland/QEUH-A638BE/2020|EPI_ISL_596159|2 ···················· ···················· ···················· ···················· ···················· ····················
M 6119 hCoV-19/Scotland/QEUH-A638DC/2020|EPI_ISL_596160|2 ···················· ···················· ···················· ···················· ···················· ····················
M 6120 hCoV-19/Scotland/QEUH-A638EB/2020|EPI_ISL_596161|2 ···················· ···················· ···················· ···················· ···················· ····················
M 6121 hCoV-19/Scotland/QEUH-A638FA/2020|EPI_ISL_596162|2 ···················· ···················· ···················· ···················· ···················· ····················
M 6122 hCoV-19/Scotland/QEUH-A63906/2020|EPI_ISL_596163|2 ···················· ···················· ···················· ···················· ···················· ····················
M 6123 hCoV-19/Scotland/QEUH-A63915/2020|EPI_ISL_596164|2 ···················· ···················· ···················· ···················· ···················· ····················
M 6124 hCoV-19/Scotland/QEUH-A63924/2020|EPI_ISL_596165|2 ···················· ···················· ···················· ···················· ···················· ····················
M 6125 hCoV-19/Scotland/QEUH-A63933/2020|EPI_ISL_596166|2 ···················· ···················· ···················· ···················· ···················· ····················
M 6126 hCoV-19/Scotland/QEUH-A63942/2020|EPI_ISL_596167|2 ···················· ···················· ···················· ···················· ···················· ····················
M 6127 hCoV-19/Scotland/QEUH-A63951/2020|EPI_ISL_596168|2 ···················· ···················· ···················· ···················· ···················· ····················
M 6128 hCoV-19/Scotland/QEUH-A63960/2020|EPI_ISL_596169|2 ···················· ···················· ···················· ···················· ···················· ····················
M 6129 hCoV-19/Scotland/QEUH-A6397F/2020|EPI_ISL_596170|2 ···················· ···················· ···················· ···················· ···················· ····················
M 6130 hCoV-19/Scotland/QEUH-A6398E/2020|EPI_ISL_596171|2 ···················· ···················· ···················· ···················· ···················· ····················
M 6131 hCoV-19/Scotland/QEUH-A6399D/2020|EPI_ISL_596172|2 ···················· ···················· ···················· ···················· ···················· ····················
M 6132 hCoV-19/Scotland/QEUH-A639BB/2020|EPI_ISL_596174|2 ···················· ···················· ···················· ···················· ···················· ····················
M 6133 hCoV-19/Scotland/QEUH-A639D9/2020|EPI_ISL_596176|2 ···················· ···················· ···················· ···················· ···················· ····················
M 6134 hCoV-19/Scotland/QEUH-A639E8/2020|EPI_ISL_596177|2 ···················· ···················· ···················· ···················· ···················· ····················
M 6135 hCoV-19/Scotland/QEUH-A639F7/2020|EPI_ISL_596178|2 ···················· ···················· ···················· ···················· ···················· ····················
M 6136 hCoV-19/Scotland/QEUH-A63AD6/2020|EPI_ISL_596179|2 ···················· ···················· ···················· ···················· ···················· ····················
M 6137 hCoV-19/Scotland/QEUH-A63B00/2020|EPI_ISL_596180|2 ···················· ···················· ···················· ···················· ···················· ····················
M 6138 hCoV-19/Scotland/QEUH-A63B1F/2020|EPI_ISL_596181|2 ···················· ···················· ···················· ···················· ···················· ····················
M 6139 hCoV-19/Scotland/QEUH-A63B2E/2020|EPI_ISL_596182|2 ···················· ···················· ···················· ···················· ···················· ····················
M 6140 hCoV-19/Scotland/QEUH-A63B3D/2020|EPI_ISL_596183|2 ···················· ···················· ···················· ···················· ···················· ····················
M 6141 hCoV-19/Scotland/QEUH-A63B4C/2020|EPI_ISL_596184|2 ···················· ···················· ···················· ···················· ···················· ····················
M 6142 hCoV-19/Scotland/QEUH-A63B5B/2020|EPI_ISL_596185|2 ···················· ···················· ···················· ···················· ···················· ····················
M 6143 hCoV-19/Scotland/QEUH-A63B97/2020|EPI_ISL_596188|2 ···················· ···················· ···················· ···················· ···················· ····················
M 6144 hCoV-19/Scotland/QEUH-A642EC/2020|EPI_ISL_596189|2 ···················· ···················· ···················· ···················· ···················· ····················
M 6145 hCoV-19/USA/DE-DHSS-F917887/2020|EPI_ISL_584013|20 ···················· ···················· ···················· ···················· ···················· ····················
M 6146 hCoV-19/USA/DE-DHSS-F918060/2020|EPI_ISL_584014|20 ···················· ···················· ···················· ···················· ···················· ····················
M 6147 hCoV-19/Scotland/CVR4470/2020|EPI_ISL_585334|2020- ···················· ···················· ···················· ···················· ···················· ····················
M 6148 hCoV-19/Scotland/CVR4471/2020|EPI_ISL_585335|2020- ···················· ···················· ···················· ···················· ···················· ····················
M 6149 hCoV-19/Scotland/CVR4472/2020|EPI_ISL_585336|2020- ···················· ···················· ···················· ···················· ···················· ····················
M 6150 hCoV-19/Scotland/CVR4473/2020|EPI_ISL_585337|2020- ···················· ···················· ···················· ···················· ···················· ····················
M 6151 hCoV-19/Scotland/CVR4474/2020|EPI_ISL_585338|2020- ···················· ···················· ···················· ···················· ···················· ····················
M 6152 hCoV-19/Scotland/CVR4475/2020|EPI_ISL_585339|2020- ···················· ···················· ···················· ···················· ···················· ····················
M 6153 hCoV-19/Scotland/CVR4476/2020|EPI_ISL_585340|2020- ···················· ···················· ···················· ···················· ···················· ····················
M 6154 hCoV-19/Scotland/CVR4477/2020|EPI_ISL_585341|2020- ···················· ···················· ···················· ···················· ···················· ····················
M 6155 hCoV-19/Scotland/CVR4479/2020|EPI_ISL_585343|2020- ···················· ···················· ···················· ···················· ···················· ····················
M 6156 hCoV-19/Scotland/CVR4483/2020|EPI_ISL_585347|2020- ···················· ···················· ···················· ···················· ···················· ····················
M 6157 hCoV-19/Scotland/CVR4485/2020|EPI_ISL_585349|2020- ···················· ···················· ···················· ···················· ···················· ····················
M 6158 hCoV-19/Scotland/CVR4493/2020|EPI_ISL_585356|2020- ···················· ···················· ···················· ···················· ···················· ····················
M 6160 hCoV-19/Scotland/CVR4496/2020|EPI_ISL_585359|2020- ···················· ···················· ···················· ···················· ···················· ····················
M 6161 hCoV-19/Scotland/CVR4498/2020|EPI_ISL_585361|2020- ···················· ···················· ···················· ···················· ···················· ····················
M 6162 hCoV-19/Scotland/CVR4499/2020|EPI_ISL_585362|2020- ···················· ···················· ···················· ···················· ···················· ····················
M 6163 hCoV-19/Scotland/CVR4500/2020|EPI_ISL_585363|2020- ···················· ···················· ···················· ···················· ···················· ····················
M 6164 hCoV-19/Scotland/CVR4501/2020|EPI_ISL_585364|2020- ···················· ···················· ···················· ···················· ···················· ····················
M 6165 hCoV-19/Scotland/CVR4502/2020|EPI_ISL_585365|2020- ···················· ···················· ···················· ···················· ···················· ····················
M 6166 hCoV-19/Scotland/CVR4504/2020|EPI_ISL_585367|2020- ···················· ···················· ···················· ···················· ···················· ····················
M 6167 hCoV-19/Scotland/CVR4506/2020|EPI_ISL_585368|2020- ···················· ···················· ···················· ···················· ···················· ····················
M 6168 hCoV-19/Scotland/CVR4507/2020|EPI_ISL_585369|2020- ···················· ···················· ···················· ···················· ···················· ····················
M 6169 hCoV-19/Scotland/CVR4508/2020|EPI_ISL_585370|2020- ···················· ···················· ···················· ···················· ···················· ····················
M 6170 hCoV-19/Scotland/CVR4510/2020|EPI_ISL_585371|2020- ···················· ···················· ···················· ···················· ···················· ····················
M 6171 hCoV-19/Scotland/CVR4511/2020|EPI_ISL_585372|2020- ···················· ···················· ···················· ···················· ···················· ····················
M 6172 hCoV-19/Scotland/CVR4512/2020|EPI_ISL_585373|2020- ···················· ···················· ···················· ···················· ···················· ····················
M 6173 hCoV-19/Scotland/CVR4513/2020|EPI_ISL_585374|2020- ···················· ···················· ···················· ···················· ···················· ····················
M 6174 hCoV-19/Scotland/CVR4514/2020|EPI_ISL_585375|2020- ···················· ···················· ···················· ···················· ···················· ····················
M 6175 hCoV-19/Scotland/CVR4515/2020|EPI_ISL_585376|2020- ···················· ···················· ···················· ···················· ···················· ····················
M 6176 hCoV-19/Scotland/CVR4516/2020|EPI_ISL_585377|2020- ···················· ···················· ···················· ···················· ···················· ····················
M 6177 hCoV-19/England/ALDP-A2E04E/2020|EPI_ISL_586665|20 ···················· ···················· ···················· ···················· ···················· ····················
M 6178 hCoV-19/England/ALDP-A2DD2A/2020|EPI_ISL_586666|20 ···················· ···················· ···················· ···················· ···················· ····················
M 6179 hCoV-19/England/ALDP-A2DAE7/2020|EPI_ISL_586667|20 ···················· ···················· ···················· ···················· ···················· ····················
M 6180 hCoV-19/England/ALDP-A2DAD8/2020|EPI_ISL_586670|20 ···················· ···················· ···················· ···················· ···················· ····················
M 6181 hCoV-19/England/ALDP-A2DF60/2020|EPI_ISL_586671|20 ···················· ···················· ···················· ···················· ···················· ····················
M 6182 hCoV-19/England/ALDP-A2E406/2020|EPI_ISL_586672|20 ···················· ···················· ···················· ···················· ···················· ····················
M 6183 hCoV-19/England/ALDP-A2D8ED/2020|EPI_ISL_586673|20 ···················· ···················· ···················· ···················· ···················· ····················
M 6184 hCoV-19/England/ALDP-A2D319/2020|EPI_ISL_586674|20 ···················· ···················· ···················· ···················· ···················· ····················
M 6185 hCoV-19/England/ALDP-A2E4CA/2020|EPI_ISL_586675|20 ···················· ···················· ···················· ···················· ···················· ····················
M 6186 hCoV-19/England/ALDP-A2D5D7/2020|EPI_ISL_586676|20 ···················· ···················· ···················· ···················· ···················· ····················
M 6187 hCoV-19/England/ALDP-A2E318/2020|EPI_ISL_586677|20 ···················· ···················· ···················· ···················· ···················· ····················
M 6188 hCoV-19/England/ALDP-A2D5B9/2020|EPI_ISL_586678|20 ···················· ···················· ···················· ···················· ···················· ····················
M 6189 hCoV-19/England/ALDP-A2D6C5/2020|EPI_ISL_586679|20 ···················· ···················· ···················· ···················· ···················· ····················
M 6190 hCoV-19/England/ALDP-A2D58C/2020|EPI_ISL_586680|20 ···················· ···················· ···················· ···················· ···················· ····················
M 6191 hCoV-19/England/ALDP-A2D935/2020|EPI_ISL_586687|20 ···················· ···················· ···················· ···················· ···················· ····················
M 6192 hCoV-19/England/ALDP-A2D759/2020|EPI_ISL_586688|20 ···················· ···················· ···················· ···················· ···················· ····················
M 6193 hCoV-19/England/ALDP-A2DFCA/2020|EPI_ISL_586689|20 ···················· ···················· ···················· ···················· ···················· ····················
M 6194 hCoV-19/England/ALDP-A2E600/2020|EPI_ISL_586690|20 ···················· ···················· ···················· ···················· ···················· ····················
M 6195 hCoV-19/England/ALDP-A2E354/2020|EPI_ISL_586691|20 ···················· ·A·················· ···················· ···················· ···················· ····················
M 6196 hCoV-19/England/ALDP-A2DF7F/2020|EPI_ISL_586692|20 ···················· ···················· ···················· ···················· ···················· ····················

| | | | | | | | |
|---|---|---|---|---|---|---|---|
| M | 6197 hCoV-19/England/ALDP-A2E837/2020\|EPI_ISL_586693\|20 | ···················· | ···················· | ···················· | ···················· | ···················· | ···················· |
| M | 6198 hCoV-19/England/ALDP-A2D4F8/2020\|EPI_ISL_586694\|20 | ···················· | ···················· | ···················· | ···················· | ···················· | ···················· |
| M | 6199 hCoV-19/England/ALDP-A2D6A7/2020\|EPI_ISL_586695\|20 | ···················· | ···················· | ···················· | ···················· | ···················· | ···················· |
| M | 6200 hCoV-19/England/ALDP-A2DCB4/2020\|EPI_ISL_586696\|20 | ···················· | ···················· | ···················· | ···················· | ···················· | ···················· |
| M | 6201 hCoV-19/England/ALDP-A2E1E1/2020\|EPI_ISL_586697\|20 | ···················· | ···················· | ···················· | ···················· | ···················· | ···················· |
| M | 6202 hCoV-19/England/ALDP-A2D786/2020\|EPI_ISL_586698\|20 | ···················· | ···················· | ···················· | ···················· | ···················· | ···················· |
| M | 6203 hCoV-19/England/ALDP-A2E11E/2020\|EPI_ISL_586699\|20 | ···················· | ···················· | ···················· | ···················· | ···················· | ···················· |
| M | 6204 hCoV-19/England/ALDP-9FCA97/2020\|EPI_ISL_587900\|20 | ···················· | ···················· | ···················· | ···················· | ···················· | ···················· |
| M | 6205 hCoV-19/England/ALDP-9FD559/2020\|EPI_ISL_587901\|20 | ···················· | ···················· | ···················· | ···················· | ···················· | ···················· |
| M | 6206 hCoV-19/England/ALDP-9FD076/2020\|EPI_ISL_587903\|20 | ···················· | ···················· | ···················· | ···················· | ···················· | ···················· |
| M | 6207 hCoV-19/England/ALDP-9FDB84/2020\|EPI_ISL_587904\|20 | ···················· | ···················· | ···················· | ···················· | ···················· | ···················· |
| M | 6208 hCoV-19/England/ALDP-9FDF00/2020\|EPI_ISL_587905\|20 | ···················· | ···················· | ···················· | ···················· | ···················· | ···················· |
| M | 6209 hCoV-19/England/ALDP-9FCF3E/2020\|EPI_ISL_587906\|20 | ···················· | ···················· | ···················· | ···················· | ···················· | ···················· |
| M | 6210 hCoV-19/England/ALDP-9FCD9E/2020\|EPI_ISL_587908\|20 | ···················· | ···················· | ···················· | ···················· | ···················· | ···················· |
| M | 6211 hCoV-19/England/ALDP-9FD2CB/2020\|EPI_ISL_587909\|20 | ···················· | ·A·······A·········· | ···················· | ···················· | ···················· | ···················· |
| M | 6212 hCoV-19/England/ALDP-9FDD15/2020\|EPI_ISL_587996\|20 | ···················· | ···················· | ···················· | ···················· | ···················· | ···················· |
| M | 6213 hCoV-19/England/ALDP-9FCB1C/2020\|EPI_ISL_587997\|20 | ···················· | ···················· | ···················· | ···················· | ···················· | ···················· |
| M | 6214 hCoV-19/England/ALDP-9FCC55/2020\|EPI_ISL_587998\|20 | ···················· | ···················· | ···················· | ···················· | ···················· | ···················· |
| M | 6215 hCoV-19/Canada/ON-S1430/2020\|EPI_ISL_591101\|2020-0 | ···················· | ···················· | ···················· | ···················· | ···················· | ···················· |
| M | 6216 hCoV-19/Canada/ON-S1468/2020\|EPI_ISL_591104\|2020-0 | ···················· | ···················· | ···················· | ···················· | ···················· | ···················· |
| M | 6217 hCoV-19/Canada/ON-S1471/2020\|EPI_ISL_591107\|2020-0 | ···················· | ···················· | ···················· | ···················· | ···················· | ···················· |
| M | 6218 hCoV-19/Canada/ON-S1472/2020\|EPI_ISL_591108\|2020-0 | ···················· | ···················· | ···················· | ···················· | ···················· | ···················· |
| M | 6219 hCoV-19/Canada/ON-S1473/2020\|EPI_ISL_591109\|2020-0 | ···················· | ···················· | ···················· | ···················· | ···················· | ···················· |
| M | 6220 hCoV-19/Canada/ON-S377/2020\|EPI_ISL_591137\|2020-03 | ···················· | ···················· | ···················· | ···················· | ···················· | ···················· |
| M | 6221 hCoV-19/Canada/ON-S707/2020\|EPI_ISL_591141\|2020-04 | ···················· | ···················· | ···················· | ···················· | ···················· | ···················· |
| M | 6222 hCoV-19/Canada/ON-S708/2020\|EPI_ISL_591142\|2020-04 | ···················· | ···················· | ···················· | ···················· | ···················· | ···················· |
| M | 6223 hCoV-19/Canada/ON-S710/2020\|EPI_ISL_591144\|2020-04 | ···················· | ···················· | ···················· | ···················· | ···················· | ···················· |
| M | 6224 hCoV-19/Canada/ON-S720/2020\|EPI_ISL_591145\|2020-04 | ···················· | ···················· | ···················· | ···················· | ···················· | ···················· |
| M | 6225 hCoV-19/Canada/ON-S722/2020\|EPI_ISL_591147\|2020-04 | ···················· | ···················· | ···················· | ···················· | ···················· | ···················· |
| M | 6226 hCoV-19/Canada/ON-S724/2020\|EPI_ISL_591148\|2020-04 | ···················· | ···················· | ···················· | ···················· | ···················· | ···················· |
| M | 6227 hCoV-19/Canada/ON-S725/2020\|EPI_ISL_591149\|2020-04 | ···················· | ···················· | ···················· | ···················· | ···················· | ···················· |
| M | 6228 hCoV-19/Canada/ON-S726/2020\|EPI_ISL_591150\|2020-04 | ···················· | ···················· | ···················· | ···················· | ···················· | ···················· |
| M | 6229 hCoV-19/Canada/ON-S727/2020\|EPI_ISL_591151\|2020-04 | ···················· | ···················· | ···················· | ···················· | ···················· | ···················· |
| M | 6230 hCoV-19/Canada/ON-S728/2020\|EPI_ISL_591152\|2020-04 | ···················· | ···················· | ···················· | ···················· | ···················· | ···················· |
| M | 6231 hCoV-19/Canada/ON-S729/2020\|EPI_ISL_591153\|2020-04 | ···················· | ···················· | ···················· | ···················· | ···················· | ···················· |
| M | 6232 hCoV-19/Canada/ON-S730/2020\|EPI_ISL_591154\|2020-04 | ···················· | ···················· | ···················· | ···················· | ···················· | ···················· |
| M | 6233 hCoV-19/Canada/ON-S732/2020\|EPI_ISL_591155\|2020-04 | ···················· | ···················· | ···················· | ···················· | ···················· | ···················· |
| M | 6234 hCoV-19/Canada/ON-S733/2020\|EPI_ISL_591156\|2020-04 | ···················· | ···················· | ···················· | ···················· | ···················· | ···················· |
| M | 6235 hCoV-19/Canada/ON-S736/2020\|EPI_ISL_591157\|2020-04 | ···················· | ···················· | ···················· | ···················· | ···················· | ···················· |
| M | 6236 hCoV-19/Canada/ON-S744/2020\|EPI_ISL_591158\|2020-04 | ···················· | ···················· | ···················· | ···················· | ···················· | ···················· |
| M | 6237 hCoV-19/Canada/ON-S754/2020\|EPI_ISL_591159\|2020-04 | ···················· | ···················· | ···················· | ···················· | ···················· | ···················· |
| M | 6238 hCoV-19/Canada/ON-S761/2020\|EPI_ISL_591160\|2020-04 | ···················· | ···················· | ···················· | ···················· | ···················· | ···················· |
| M | 6239 hCoV-19/Canada/ON-S797/2020\|EPI_ISL_591161\|2020-04 | ···················· | ···················· | ···················· | ···················· | ···················· | ···················· |
| M | 6240 hCoV-19/Canada/ON-S803/2020\|EPI_ISL_591162\|2020-04 | ···················· | ···················· | ···················· | ···················· | ···················· | ···················· |
| M | 6241 hCoV-19/Canada/ON-S804/2020\|EPI_ISL_591163\|2020-04 | ···················· | ···················· | ···················· | ···················· | ···················· | ···················· |
| M | 6242 hCoV-19/Canada/ON-S811/2020\|EPI_ISL_591164\|2020-04 | ···················· | ···················· | ···················· | ···················· | ···················· | ···················· |
| M | 6243 hCoV-19/Canada/ON-S813/2020\|EPI_ISL_591165\|2020-04 | ···················· | ···················· | ···················· | ···················· | ···················· | ···················· |
| M | 6244 hCoV-19/Canada/ON-S815/2020\|EPI_ISL_591166\|2020-04 | ···················· | ···················· | ···················· | ···················· | ···················· | ···················· |
| M | 6245 hCoV-19/Canada/ON-S816/2020\|EPI_ISL_591167\|2020-04 | ···················· | ···················· | ···················· | ···················· | ···················· | ···················· |
| M | 6246 hCoV-19/Canada/ON-S846/2020\|EPI_ISL_591169\|2020-04 | ···················· | ···················· | ···················· | ···················· | ···················· | ···················· |
| M | 6247 hCoV-19/Canada/ON-S851/2020\|EPI_ISL_591170\|2020-04 | ···················· | ···················· | ···················· | ···················· | ······A············· | ···················· |
| M | 6248 hCoV-19/Canada/ON-S857/2020\|EPI_ISL_591171\|2020-04 | ···················· | ···················· | ···················· | ···················· | ···················· | ···················· |
| M | 6249 hCoV-19/Canada/ON-S859/2020\|EPI_ISL_591172\|2020-04 | ···················· | ···················· | ···················· | ···················· | ···················· | ···················· |
| M | 6250 hCoV-19/Canada/ON-S870/2020\|EPI_ISL_591173\|2020-04 | ···················· | ···················· | ···················· | ···················· | ···················· | ···················· |
| M | 6251 hCoV-19/Canada/ON-S873/2020\|EPI_ISL_591174\|2020-04 | ···················· | ···················· | ···················· | ···················· | ···················· | ···················· |
| M | 6252 hCoV-19/Australia/VIC14224/2020\|EPI_ISL_592400\|202 | ···················· | ···················· | ···················· | ···················· | ···················· | ···················· |
| M | 6253 hCoV-19/Australia/VIC14225/2020\|EPI_ISL_592401\|202 | ···················· | ···················· | ···················· | ···················· | ···················· | ···················· |
| M | 6254 hCoV-19/Australia/VIC14227/2020\|EPI_ISL_592402\|202 | ···················· | ···················· | ···················· | ···················· | ···················· | ···················· |
| M | 6255 hCoV-19/Australia/VIC14230/2020\|EPI_ISL_592403\|202 | ···················· | ···················· | ···················· | ···················· | ···················· | ···················· |
| M | 6256 hCoV-19/Australia/VIC14231/2020\|EPI_ISL_592404\|202 | ···················· | ···················· | ···················· | ···················· | ···················· | ···················· |
| M | 6257 hCoV-19/Australia/VIC14232/2020\|EPI_ISL_592405\|202 | ···················· | ···················· | ···················· | ···················· | ···················· | ···················· |
| M | 6258 hCoV-19/Australia/VIC14234/2020\|EPI_ISL_592406\|202 | ···················· | ···················· | ···················· | ···················· | ···················· | ···················· |
| M | 6259 hCoV-19/Australia/VIC14235/2020\|EPI_ISL_592407\|202 | ···················· | ···················· | ···················· | ···················· | ···················· | ···················· |
| M | 6260 hCoV-19/Australia/VIC14238/2020\|EPI_ISL_592408\|202 | ···················· | ···················· | ···················· | ···················· | ···················· | ···················· |
| M | 6261 hCoV-19/Australia/VIC14239/2020\|EPI_ISL_592409\|202 | ···················· | ···················· | ···················· | ···················· | ···················· | ···················· |
| M | 6262 hCoV-19/Australia/VIC14241/2020\|EPI_ISL_592410\|202 | ···················· | ···················· | ···················· | ···················· | ···················· | ···················· |
| M | 6263 hCoV-19/Australia/VIC14243/2020\|EPI_ISL_592411\|202 | ···················· | ···················· | ···················· | ···················· | ···················· | ···················· |
| M | 6264 hCoV-19/Australia/VIC14244/2020\|EPI_ISL_592412\|202 | ···················· | ···················· | ···················· | ···················· | ···················· | ···················· |
| M | 6265 hCoV-19/Australia/VIC14245/2020\|EPI_ISL_592413\|202 | ···················· | ···················· | ···················· | ···················· | ···················· | ···················· |
| M | 6266 hCoV-19/Australia/VIC14246/2020\|EPI_ISL_592414\|202 | ···················· | ···················· | ···················· | ···················· | ···················· | ···················· |
| M | 6267 hCoV-19/Australia/VIC14247/2020\|EPI_ISL_592415\|202 | ···················· | ···················· | ···················· | ···················· | ···················· | ···················· |
| M | 6268 hCoV-19/Australia/VIC14248/2020\|EPI_ISL_592416\|202 | ···················· | ···················· | ···················· | ···················· | ···················· | ···················· |
| M | 6269 hCoV-19/Australia/VIC14249/2020\|EPI_ISL_592417\|202 | ···················· | ···················· | ···················· | ···················· | ···················· | ···················· |
| M | 6270 hCoV-19/Australia/VIC14251/2020\|EPI_ISL_592418\|202 | ···················· | ···················· | ···················· | ···················· | ···················· | ···················· |
| M | 6271 hCoV-19/Australia/VIC14254/2020\|EPI_ISL_592419\|202 | ···················· | ···················· | ···················· | ···················· | N··················· | ···················· |
| M | 6272 hCoV-19/Australia/VIC14279/2020\|EPI_ISL_592430\|202 | ···················· | ···················· | ···················· | ···················· | ···················· | ···················· |
| M | 6273 hCoV-19/Australia/VIC14280/2020\|EPI_ISL_592431\|202 | ···················· | ···················· | ···················· | ···················· | ···················· | ···················· |
| M | 6274 hCoV-19/Australia/VIC14282/2020\|EPI_ISL_592432\|202 | ···················· | ···················· | ···················· | ···················· | ···················· | ···················· |
| M | 6275 hCoV-19/Australia/VIC14283/2020\|EPI_ISL_592433\|202 | ···················· | ···················· | ···················· | ···················· | ···················· | ···················· |
| M | 6276 hCoV-19/Australia/VIC14284/2020\|EPI_ISL_592434\|202 | ···················· | ···················· | ···················· | ···················· | ···················· | ···················· |

| | | | | | | | |
|---|---|---|---|---|---|---|---|
| M | 6277 hCoV-19/Australia/VIC14287/2020\|EPI_ISL_592436\|202 | ···················· | ···················· | ····················· | ···················· | ···················· | ···················· |
| M | 6278 hCoV-19/Australia/VIC14290/2020\|EPI_ISL_592437\|202 | ···················· | ···················· | ····················· | ···················· | ···················· | ···················· |
| M | 6279 hCoV-19/Australia/VIC14292/2020\|EPI_ISL_592438\|202 | ···················· | ···················· | ····················· | ···················· | ···················· | ···················· |
| M | 6280 hCoV-19/Australia/VIC14294/2020\|EPI_ISL_592439\|202 | ···················· | ···················· | ····················· | ···················· | ···················· | ···················· |
| M | 6281 hCoV-19/Australia/VIC14350/2020\|EPI_ISL_592462\|202 | ···················· | ···················· | ····················· | ···················· | ···················· | ···················· |
| M | 6282 hCoV-19/Australia/VIC14352/2020\|EPI_ISL_592463\|202 | ···················· | ···················· | ····················· | ···················· | ···················· | ···················· |
| M | 6283 hCoV-19/Australia/VIC14353/2020\|EPI_ISL_592464\|202 | ···················· | ···················· | ····················· | ···················· | ···················· | ···················· |
| M | 6284 hCoV-19/Australia/VIC14354/2020\|EPI_ISL_592465\|202 | ···················· | ···················· | ····················· | ···················· | ···················· | ···················· |
| M | 6286 hCoV-19/Australia/VIC14359/2020\|EPI_ISL_592467\|202 | ···················· | ···················· | ····················· | ···················· | ···················· | ···················· |
| M | 6287 hCoV-19/Australia/VIC14365/2020\|EPI_ISL_592468\|202 | ···················· | ···················· | ····················· | ···················· | ···················· | ···················· |
| M | 6288 hCoV-19/Australia/VIC14367/2020\|EPI_ISL_592469\|202 | ···················· | ···················· | ····················· | ···················· | ···················· | ···················· |
| M | 6289 hCoV-19/Australia/VIC14368/2020\|EPI_ISL_592470\|202 | ···················· | ···················· | ····················· | ···················· | ···················· | ···················· |
| M | 6290 hCoV-19/Australia/VIC14369/2020\|EPI_ISL_592471\|202 | ···················· | ···················· | ····················· | ···················· | ···················· | ···················· |
| M | 6291 hCoV-19/Australia/VIC14372/2020\|EPI_ISL_592472\|202 | ···················· | ···················· | ····················· | ···················· | ···················· | ···················· |
| M | 6293 hCoV-19/Australia/VIC14376/2020\|EPI_ISL_592474\|202 | ···················· | ···················· | ····················· | ···················· | ···················· | ···················· |
| M | 6295 hCoV-19/Australia/VIC14378/2020\|EPI_ISL_592476\|202 | ···················· | ···················· | ····················· | ···················· | ···················· | ···················· |
| M | 6296 hCoV-19/Australia/VIC14380/2020\|EPI_ISL_592477\|202 | ···················· | ···················· | ····················· | ···················· | ···················· | ···················· |
| M | 6297 hCoV-19/Australia/VIC14381/2020\|EPI_ISL_592478\|202 | ···················· | ···················· | ····················· | ···················· | ···················· | ···················· |
| M | 6298 hCoV-19/Australia/VIC14385/2020\|EPI_ISL_592479\|202 | ···················· | ···················· | ····················· | ···················· | ···················· | ···················· |
| M | 6299 hCoV-19/Australia/VIC14387/2020\|EPI_ISL_592480\|202 | ···················· | ···················· | ····················· | ···················· | ···················· | ···················· |
| M | 6302 hCoV-19/Australia/VIC14398/2020\|EPI_ISL_592483\|202 | ···················· | ···················· | ····················· | ···················· | ···················· | ···················· |
| M | 6304 hCoV-19/Australia/VIC14406/2020\|EPI_ISL_592485\|202 | ···················· | ···················· | ··K·················· | ···················· | ···················· | ···················· |
| M | 6305 hCoV-19/Australia/VIC14407/2020\|EPI_ISL_592486\|202 | ···················· | ···················· | ····················· | ···················· | ···················· | ···················· |
| M | 6306 hCoV-19/Australia/VIC14408/2020\|EPI_ISL_592487\|202 | ···················· | ···················· | ····················· | ···················· | ···················· | ···················· |
| M | 6307 hCoV-19/Australia/VIC14410/2020\|EPI_ISL_592488\|202 | ···················· | ···················· | ····················· | ···················· | ···················· | ···················· |
| M | 6308 hCoV-19/Australia/VIC14412/2020\|EPI_ISL_592489\|202 | ···················· | ···················· | ····················· | ···················· | ···················· | ···················· |
| M | 6309 hCoV-19/Australia/VIC14414/2020\|EPI_ISL_592490\|202 | ···················· | ···················· | ····················· | ···················· | ······K············· | ···················· |
| M | 6310 hCoV-19/Australia/VIC14415/2020\|EPI_ISL_592491\|202 | ···················· | ···················· | ····················· | ···················· | ···················· | ···················· |
| M | 6311 hCoV-19/Australia/VIC14418/2020\|EPI_ISL_592492\|202 | ···················· | ···················· | ····················· | ···················· | ···················· | ···················· |
| M | 6312 hCoV-19/Australia/VIC14419/2020\|EPI_ISL_592493\|202 | ···················· | ···················· | ····················· | ···················· | ···················· | ···················· |
| M | 6313 hCoV-19/Australia/VIC14420/2020\|EPI_ISL_592494\|202 | ···················· | ···················· | ····················· | ···················· | ···················· | ···················· |
| M | 6314 hCoV-19/Australia/VIC14421/2020\|EPI_ISL_592495\|202 | ···················· | ···················· | ····················· | ···················· | ···R················ | ···················· |
| M | 6316 hCoV-19/Australia/VIC14424/2020\|EPI_ISL_592497\|202 | ···················· | ···················· | ····················· | ···················· | ···················· | ···················· |
| M | 6317 hCoV-19/Australia/VIC14426/2020\|EPI_ISL_592498\|202 | ···················· | ···················· | ····················· | ···················· | ···················· | ···················· |
| M | 6319 hCoV-19/Australia/NSW1048/2020\|EPI_ISL_593700\|2020 | ···················· | ·A·················· | ····················· | ···················· | ···················· | ··A················· |
| M | 6320 hCoV-19/Australia/NSW1049/2020\|EPI_ISL_593701\|2020 | ···················· | ·A·················· | ····················· | ···················· | ···················· | ··A················· |
| M | 6321 hCoV-19/Australia/NSW1050/2020\|EPI_ISL_593702\|2020 | ···················· | ···················· | ····················· | ···················· | ···················· | ···················· |
| M | 6322 hCoV-19/Australia/NSW1051/2020\|EPI_ISL_593703\|2020 | ···················· | ···················· | ····················· | ···················· | ···················· | ···················· |
| M | 6323 hCoV-19/Australia/NSW1052/2020\|EPI_ISL_593704\|2020 | ···················· | ·A·················· | ····················· | ···················· | ···················· | ···················· |
| M | 6324 hCoV-19/Australia/NSW1053/2020\|EPI_ISL_593705\|2020 | ···················· | ···················· | ····················· | ···················· | ···················· | ···················· |
| M | 6325 hCoV-19/Australia/NSW1054/2020\|EPI_ISL_593706\|2020 | ···················· | ···················· | ····················· | ···················· | ···················· | ···················· |
| M | 6326 hCoV-19/Australia/NSW1055/2020\|EPI_ISL_593707\|2020 | ···················· | ·A·················· | ····················· | ···················· | ···················· | ··A················· |
| M | 6327 hCoV-19/Australia/NSW1068/2020\|EPI_ISL_593708\|2020 | ···················· | ···················· | ····················· | ···················· | ···················· | ···················· |
| M | 6328 hCoV-19/Australia/NSW1069/2020\|EPI_ISL_593709\|2020 | ···················· | ···················· | ····················· | ···················· | ···················· | ···················· |
| M | 6329 hCoV-19/Australia/NSW1076/2020\|EPI_ISL_593710\|2020 | ···················· | ···················· | ····················· | ···················· | ···················· | ···················· |
| M | 6330 hCoV-19/Australia/NSW1077/2020\|EPI_ISL_593711\|2020 | ···················· | ···················· | ····················· | ···················· | ···················· | ···················· |
| M | 6331 hCoV-19/Australia/NSW1083/2020\|EPI_ISL_593712\|2020 | ···················· | ···················· | ····················· | ···················· | ···················· | ···················· |
| M | 6332 hCoV-19/Australia/NSW1098/2020\|EPI_ISL_593713\|2020 | ···················· | ·A·················· | ····················· | ···················· | ···················· | ···················· |
| M | 6333 hCoV-19/Australia/NSW1099/2020\|EPI_ISL_593714\|2020 | ···················· | ···················· | ····················· | ···················· | ···················· | ···················· |
| M | 6334 hCoV-19/Australia/NSW1100/2020\|EPI_ISL_593715\|2020 | ···················· | ·A·················· | ····················· | ···················· | ···················· | ··A················· |
| M | 6335 hCoV-19/Australia/NSW1101/2020\|EPI_ISL_593716\|2020 | ···················· | ·A·················· | ····················· | ···················· | ···················· | ··A················· |
| M | 6336 hCoV-19/Australia/NSW1102/2020\|EPI_ISL_593717\|2020 | ···················· | ·A·················· | ····················· | ···················· | ···················· | ··A················· |
| M | 6337 hCoV-19/Australia/NSW1103/2020\|EPI_ISL_593718\|2020 | ···················· | ···················· | ····················· | ···················· | ···················· | ···················· |
| M | 6338 hCoV-19/Australia/NSW1104/2020\|EPI_ISL_593719\|2020 | ···················· | ···················· | ····················· | ···················· | ···················· | ···················· |
| M | 6339 hCoV-19/Australia/NSW1105/2020\|EPI_ISL_593720\|2020 | ···················· | ···················· | ····················· | ···················· | ···················· | ···················· |
| M | 6340 hCoV-19/Australia/NSW1106/2020\|EPI_ISL_593721\|2020 | ···················· | ···················· | ····················· | ···················· | ···················· | ···················· |
| M | 6341 hCoV-19/Australia/NSW1107/2020\|EPI_ISL_593722\|2020 | ···················· | ···················· | ····················· | ···················· | ···················· | ···················· |
| M | 6342 hCoV-19/Australia/NSW1108/2020\|EPI_ISL_593723\|2020 | ···················· | ···················· | ····················· | ···················· | ···················· | ···················· |
| M | 6343 hCoV-19/Australia/NSW1109/2020\|EPI_ISL_593724\|2020 | ···················· | ·A·················· | ····················· | ···················· | ···················· | ···················· |
| M | 6344 hCoV-19/Australia/NSW1111/2020\|EPI_ISL_593726\|2020 | ···················· | ···················· | ····················· | ···················· | ···················· | ···················· |
| M | 6345 hCoV-19/Australia/NSW1112/2020\|EPI_ISL_593727\|2020 | ···················· | ···················· | ····················· | ···················· | ···················· | ···················· |
| M | 6346 hCoV-19/Australia/NSW1113/2020\|EPI_ISL_593728\|2020 | ···················· | ···················· | ····················· | ···················· | ···················· | ···················· |
| M | 6347 hCoV-19/Australia/NSW1115/2020\|EPI_ISL_593729\|2020 | ···················· | ···················· | ····················· | ···················· | ···················· | ···················· |
| M | 6348 hCoV-19/Australia/NSW1116/2020\|EPI_ISL_593730\|2020 | ···················· | ···················· | ····················· | ···················· | ···················· | ···················· |
| M | 6349 hCoV-19/Australia/NSW1117/2020\|EPI_ISL_593731\|2020 | ···················· | ·A·················· | ····················· | ···················· | ···················· | ··A················· |
| M | 6350 hCoV-19/Australia/NSW1118/2020\|EPI_ISL_593732\|2020 | ···················· | ···················· | ····················· | ···················· | ···················· | ···················· |
| M | 6351 hCoV-19/Australia/NSW1119/2020\|EPI_ISL_593733\|2020 | ···················· | ···················· | ····················· | ···················· | ···················· | ···················· |
| M | 6352 hCoV-19/Australia/NSW1120/2020\|EPI_ISL_593734\|2020 | ···················· | ·A·················· | ····················· | ···················· | ···················· | ··A················· |
| M | 6353 hCoV-19/Australia/NSW1121/2020\|EPI_ISL_593735\|2020 | ···················· | ···················· | ····················· | ···················· | ···················· | ···················· |
| M | 6354 hCoV-19/Australia/NSW1122/2020\|EPI_ISL_593736\|2020 | ···················· | ···················· | ····················· | ···················· | ···················· | ···················· |
| M | 6355 hCoV-19/Australia/NSW1123/2020\|EPI_ISL_593737\|2020 | ···················· | ···················· | ····················· | ···················· | ···················· | ···················· |
| M | 6356 hCoV-19/Australia/NSW1124/2020\|EPI_ISL_593738\|2020 | ···················· | ···················· | ····················· | ···················· | ···················· | ···················· |
| M | 6357 hCoV-19/Australia/NSW1125/2020\|EPI_ISL_593739\|2020 | ···················· | ···················· | ····················· | ···················· | ···················· | ···················· |
| M | 6358 hCoV-19/Australia/NSW1126/2020\|EPI_ISL_593740\|2020 | ···················· | ·A·················· | ····················· | ···················· | ···················· | ··A················· |
| M | 6359 hCoV-19/Australia/NSW1127/2020\|EPI_ISL_593741\|2020 | ···················· | ···················· | ····················· | ···················· | ···················· | ···················· |
| M | 6360 hCoV-19/Australia/NSW1129/2020\|EPI_ISL_593742\|2020 | ···················· | ···················· | ····················· | ···················· | ···················· | ···················· |
| M | 6361 hCoV-19/Australia/NSW1130/2020\|EPI_ISL_593743\|2020 | ···················· | ···················· | ····················· | ···················· | ···················· | ···················· |
| M | 6362 hCoV-19/Australia/NSW1132/2020\|EPI_ISL_593744\|2020 | ···················· | ···················· | ····················· | ···················· | ···················· | ···················· |
| M | 6363 hCoV-19/Australia/NSW1133/2020\|EPI_ISL_593745\|2020 | ···················· | ···A················ | ····················· | ···················· | ···················· | ···················· |
| M | 6364 hCoV-19/Australia/NSW1135/2020\|EPI_ISL_593746\|2020 | ···················· | ···················· | ····················· | ···················· | ···················· | ···················· |

| | | | | | | | |
|---|---|---|---|---|---|---|---|
| M | 6365 hCoV-19/Australia/NSW1136/2020\|EPI_ISL_593747\|2020 | ···················· | ···················· | ···················· | ···················· | ···················· | ···················· |
| M | 6366 hCoV-19/Australia/NSW1137/2020\|EPI_ISL_593748\|2020 | ···················· | ···················· | ···················· | ···················· | ···················· | ···················· |
| M | 6367 hCoV-19/Australia/NSW1139/2020\|EPI_ISL_593749\|2020 | ···················· | ···················· | ···················· | ···················· | ···················· | ···················· |
| M | 6368 hCoV-19/Australia/NSW1064/2020\|EPI_ISL_593760\|2020 | ···················· | ···················· | ···················· | ···················· | ···················· | ···················· |
| M | 6369 hCoV-19/Australia/NSW1073/2020\|EPI_ISL_593761\|2020 | ···················· | ···················· | ···················· | ···················· | ···················· | ···················· |
| M | 6370 hCoV-19/Australia/NSW1090/2020\|EPI_ISL_593762\|2020 | ···················· | ···················· | ···················· | ···················· | ···················· | ···················· |
| M | 6371 hCoV-19/Australia/NSW1094/2020\|EPI_ISL_593763\|2020 | ···················· | ···················· | ···················· | ···················· | ···················· | ···················· |
| M | 6372 hCoV-19/Australia/NSW1097/2020\|EPI_ISL_593764\|2020 | ···················· | ···················· | ···················· | ···················· | ···················· | ···················· |
| M | 6373 hCoV-19/Australia/NSW1131/2020\|EPI_ISL_593765\|2020 | ···················· | ···················· | ···················· | ···················· | ···················· | ···················· |
| M | 6374 hCoV-19/Australia/NSW1089/2020\|EPI_ISL_593768\|2020 | ···················· | ···················· | ···················· | ···················· | ···················· | ···················· |
| M | 6375 hCoV-19/Australia/NSW1081/2020\|EPI_ISL_593769\|2020 | ···················· | ···················· | ···················· | ···················· | ···················· | ···················· |
| M | 6376 hCoV-19/England/204221398/2020\|EPI_ISL_593793\|2020 | ···················· | ···················· | ···················· | ···················· | ···················· | ···················· |
| M | 6377 hCoV-19/England/204221399/2020\|EPI_ISL_593794\|2020 | ···················· | ···················· | ···················· | ···················· | ···················· | ···················· |
| M | 6378 hCoV-19/England/204221402/2020\|EPI_ISL_593796\|2020 | ···················· | ···················· | ···················· | ···················· | ···················· | ···················· |
| M | 6379 hCoV-19/England/204221403/2020\|EPI_ISL_593797\|2020 | ···················· | ···················· | ···················· | ···················· | ···················· | ···················· |
| M | 6380 hCoV-19/England/204221405/2020\|EPI_ISL_593798\|2020 | ···················· | ···················· | ···················· | ···················· | ···················· | ···················· |
| M | 6381 hCoV-19/Scotland/QEUH-A4C7EB/2020\|EPI_ISL_596001\|2 | ···················· | ···················· | ···················· | ···················· | ···················· | ···················· |
| M | 6382 hCoV-19/Scotland/QEUH-A4C7FA/2020\|EPI_ISL_596002\|2 | ···················· | ···················· | ···················· | ···················· | ···················· | ···················· |
| M | 6383 hCoV-19/Scotland/QEUH-A4C806/2020\|EPI_ISL_596003\|2 | ·······T············ | ···················· | ···················· | ···················· | ···················· | ···················· |
| M | 6385 hCoV-19/Scotland/QEUH-A4C833/2020\|EPI_ISL_596005\|2 | ···················· | ···················· | ···················· | ···················· | ···················· | ···················· |
| M | 6386 hCoV-19/Scotland/QEUH-A4C842/2020\|EPI_ISL_596006\|2 | ···················· | ···················· | ···················· | ···················· | ···················· | ···················· |
| M | 6387 hCoV-19/Scotland/QEUH-A4C851/2020\|EPI_ISL_596007\|2 | ···················· | ···················· | ···················· | ···················· | ···················· | ···················· |
| M | 6389 hCoV-19/Scotland/QEUH-A4C88E/2020\|EPI_ISL_596009\|2 | ···················· | ···················· | ···················· | ···················· | ···················· | ···················· |
| M | 6390 hCoV-19/Scotland/QEUH-A4CA3D/2020\|EPI_ISL_596010\|2 | ···················· | ···················· | ···················· | ···················· | ···················· | ···················· |
| M | 6391 hCoV-19/Scotland/QEUH-A4CA4C/2020\|EPI_ISL_596011\|2 | ···················· | ···················· | ···················· | ···················· | ···················· | ···················· |
| M | 6392 hCoV-19/England/QEUH-A4CA97/2020\|EPI_ISL_596014\|20 | ···················· | ···················· | ···················· | ···················· | ···················· | ···················· |
| M | 6393 hCoV-19/England/QEUH-A4CAB5/2020\|EPI_ISL_596016\|20 | ···················· | ···················· | ···················· | ···················· | ···················· | ···················· |
| M | 6394 hCoV-19/England/QEUH-A4CAC4/2020\|EPI_ISL_596017\|20 | ···················· | ···················· | ···················· | ···················· | ···················· | ···················· |
| M | 6395 hCoV-19/England/QEUH-A4CAE2/2020\|EPI_ISL_596018\|20 | ···················· | ···················· | ···················· | ···················· | ···················· | ···················· |
| M | 6396 hCoV-19/England/QEUH-A4CB0D/2020\|EPI_ISL_596020\|20 | ···················· | ···················· | ···················· | ···················· | ···················· | ···················· |
| M | 6399 hCoV-19/England/QEUH-A4CB49/2020\|EPI_ISL_596023\|20 | ···················· | ···················· | ···················· | ···················· | ···················· | ···················· |
| M | 6400 hCoV-19/England/QEUH-A4CB58/2020\|EPI_ISL_596024\|20 | ···················· | ···················· | ···················· | ···················· | ···················· | ···················· |
| M | 6401 hCoV-19/Italy/LOM-UniMI-ICCS1/2020\|EPI_ISL_584048\| | ···················· | ···················· | ···················· | ···················· | ···················· | ···················· |
| M | 6402 hCoV-19/Italy/LOM-UniMI-ICCS2/2020\|EPI_ISL_584049\| | ···················· | ···················· | ···················· | ···················· | ···················· | ···················· |
| M | 6403 hCoV-19/Italy/LOM-UniMI-ICCS3/2020\|EPI_ISL_584052\| | ···················· | ···················· | ···················· | ···················· | ···················· | ···················· |
| M | 6404 hCoV-19/England/20422146504/2020\|EPI_ISL_584053\|20 | ···················· | ···················· | ···················· | ···················· | ···················· | ···················· |
| M | 6405 hCoV-19/England/20422146604/2020\|EPI_ISL_584054\|20 | ···················· | ···················· | ···················· | ···················· | ···················· | ···················· |
| M | 6406 hCoV-19/England/20422146704/2020\|EPI_ISL_584055\|20 | ···················· | ···················· | ···················· | ···················· | ···················· | ···················· |
| M | 6407 hCoV-19/England/20422146804/2020\|EPI_ISL_584056\|20 | ···················· | ···················· | ···················· | ···················· | ···················· | ···················· |
| M | 6408 hCoV-19/England/20422146904/2020\|EPI_ISL_584057\|20 | ···················· | ···················· | ···················· | ···················· | ···················· | ···················· |
| M | 6409 hCoV-19/England/20422147004/2020\|EPI_ISL_584058\|20 | ···················· | ···················· | ···················· | ···················· | ···················· | ···················· |
| M | 6410 hCoV-19/England/20422147204/2020\|EPI_ISL_584060\|20 | ···················· | ···················· | ···················· | ···················· | ···················· | ···················· |
| M | 6411 hCoV-19/England/20422147304/2020\|EPI_ISL_584061\|20 | ···················· | ···················· | ···················· | ···················· | ···················· | ···················· |
| M | 6412 hCoV-19/England/20422147404/2020\|EPI_ISL_584062\|20 | ···················· | ···················· | ···················· | ···················· | ···················· | ···················· |
| M | 6413 hCoV-19/England/20422147504/2020\|EPI_ISL_584063\|20 | ···················· | ···················· | ···················· | ···················· | ···················· | ···················· |
| M | 6414 hCoV-19/England/20422147704/2020\|EPI_ISL_584064\|20 | ···················· | ···················· | ···················· | ···················· | ···················· | ···················· |
| M | 6415 hCoV-19/England/20422147804/2020\|EPI_ISL_584065\|20 | ···················· | ···················· | ···················· | ···················· | ···················· | ···················· |
| M | 6416 hCoV-19/England/20422148204/2020\|EPI_ISL_584066\|20 | ···················· | ···················· | ···················· | ···················· | ···················· | ···················· |
| M | 6417 hCoV-19/England/20422148504/2020\|EPI_ISL_584068\|20 | ···················· | ···················· | ···················· | ···················· | ···················· | ···················· |
| M | 6418 hCoV-19/Italy/CAM-IZSM-43083/2020\|EPI_ISL_584069\|2 | ···················· | ···················· | ···················· | ···················· | ···················· | ···················· |
| M | 6419 hCoV-19/Italy/CAM-IZSM-45947/2020\|EPI_ISL_584071\|2 | ···················· | ···················· | ···················· | ···················· | ···················· | ···················· |
| M | 6420 hCoV-19/Italy/CAM-IZSM-183/2020\|EPI_ISL_584072\|202 | ···················· | ···················· | ···················· | ···················· | ···················· | ···················· |
| M | 6421 hCoV-19/Czech Republic/NRL-6847-2/2020\|EPI_ISL_584 | ···················· | ···················· | ···················· | ···················· | ···················· | ···················· |
| M | 6422 hCoV-19/Czech Republic/NRL-7322-1/2020\|EPI_ISL_584 | ···················· | ···················· | ···················· | ···················· | ···················· | ···················· |
| M | 6423 hCoV-19/Czech Republic/NRL-7322-2/2020\|EPI_ISL_584 | ···················· | ···················· | ···················· | ···················· | ···················· | ···················· |
| M | 6424 hCoV-19/Czech Republic/NRL-7322-3/2020\|EPI_ISL_584 | ···················· | ···················· | ···················· | ···················· | ···················· | ···················· |
| M | 6425 hCoV-19/Czech Republic/NRL-7322-4/2020\|EPI_ISL_584 | ···················· | ···················· | ···················· | ···················· | ···················· | ···················· |
| M | 6426 hCoV-19/Czech Republic/NRL-7963/2020\|EPI_ISL_58407 | ···················· | ···················· | ···················· | ···················· | ···················· | ···················· |
| M | 6427 hCoV-19/Czech Republic/NRL-5593/2020\|EPI_ISL_58407 | ···················· | ···················· | ···················· | ···················· | ···················· | ···················· |
| M | 6428 hCoV-19/Czech Republic/NRL-6781/2020\|EPI_ISL_58408 | ···················· | ···················· | ···················· | ···················· | ···················· | ···················· |
| M | 6429 hCoV-19/Czech Republic/NRL-8598/2020\|EPI_ISL_58408 | ···················· | ···················· | ···················· | ···················· | ···················· | ···················· |
| M | 6430 hCoV-19/Czech Republic/NRL-8609/2020\|EPI_ISL_58408 | ···················· | ······C············· | ···················· | ···················· | ···················· | ···················· |
| M | 6431 hCoV-19/Scotland/CVR4429/2020\|EPI_ISL_585300\|2020- | ···················· | ···················· | ···················· | ···················· | ···················· | ···················· |
| M | 6432 hCoV-19/Scotland/CVR4433/2020\|EPI_ISL_585303\|2020- | ···················· | ···················· | ···················· | ···················· | ···················· | ···················· |
| M | 6433 hCoV-19/Scotland/CVR4434/2020\|EPI_ISL_585304\|2020- | ···················· | ···················· | ···················· | ···················· | ···················· | ···················· |
| M | 6434 hCoV-19/Scotland/CVR4435/2020\|EPI_ISL_585305\|2020- | ···················· | ···················· | ···················· | ···················· | ···················· | ···················· |
| M | 6435 hCoV-19/Scotland/CVR4436/2020\|EPI_ISL_585306\|2020- | ···················· | ···················· | ···················· | ···················· | ···················· | ···················· |
| M | 6436 hCoV-19/Scotland/CVR4437/2020\|EPI_ISL_585307\|2020- | ···················· | ···················· | ···················· | ···················· | ···················· | ···················· |
| M | 6437 hCoV-19/Scotland/CVR4441/2020\|EPI_ISL_585310\|2020- | ···················· | ···················· | ···················· | ···················· | ···················· | ···················· |
| M | 6439 hCoV-19/Scotland/CVR4443/2020\|EPI_ISL_585312\|2020- | ···················· | ···················· | ···················· | ···················· | ···················· | ···················· |
| M | 6440 hCoV-19/Scotland/CVR4444/2020\|EPI_ISL_585313\|2020- | ···················· | ···················· | ···················· | ···················· | ···················· | ···················· |
| M | 6441 hCoV-19/Scotland/CVR4448/2020\|EPI_ISL_585315\|2020- | ···················· | ···················· | ···················· | ···················· | ···················· | ···················· |
| M | 6442 hCoV-19/Scotland/CVR4449/2020\|EPI_ISL_585316\|2020- | ···················· | ···················· | ···················· | ···················· | ···················· | ···················· |
| M | 6443 hCoV-19/Scotland/CVR4450/2020\|EPI_ISL_585317\|2020- | ···················· | ···················· | ···················· | ···················· | ···················· | ···················· |
| M | 6444 hCoV-19/Scotland/CVR4451/2020\|EPI_ISL_585318\|2020- | ···················· | ···················· | ···················· | ···················· | ···················· | ···················· |
| M | 6445 hCoV-19/Scotland/CVR4454/2020\|EPI_ISL_585319\|2020- | ···················· | ···················· | ···················· | ···················· | ···················· | ···N··NN············ 1 |
| M | 6446 hCoV-19/Scotland/CVR4455/2020\|EPI_ISL_585320\|2020- | ···················· | ···················· | ···················· | ···················· | ···················· | ···················· |
| M | 6447 hCoV-19/Scotland/CVR4456/2020\|EPI_ISL_585321\|2020- | ···················· | ···················· | ···················· | ···················· | ···················· | ···················· |
| M | 6448 hCoV-19/Scotland/CVR4457/2020\|EPI_ISL_585322\|2020- | ···················· | ···················· | ···················· | ···················· | ···················· | ···N················ |
| M | 6449 hCoV-19/Scotland/CVR4458/2020\|EPI_ISL_585323\|2020- | ···················· | ···················· | ···················· | ···················· | ···················· | ···················· |

| | | | | | | | |
|---|---|---|---|---|---|---|---|
| M | 6450 hCoV-19/Scotland/CVR4459/2020\|EPI_ISL_585324\|2020- | ···················· | ···················· | ···················· | ···················· | ···················· | ···················· |
| M | 6451 hCoV-19/Scotland/CVR4460/2020\|EPI_ISL_585325\|2020- | ···················· | ···················· | ···················· | ···················· | ···················· | ···················· |
| M | 6452 hCoV-19/Scotland/CVR4461/2020\|EPI_ISL_585326\|2020- | ···················· | ···················· | ···················· | ···················· | ···················· | ···················· |
| M | 6453 hCoV-19/Scotland/CVR4463/2020\|EPI_ISL_585328\|2020- | ···················· | ···················· | ···················· | ···················· | ···················· | ···················· |
| M | 6454 hCoV-19/Scotland/CVR4465/2020\|EPI_ISL_585329\|2020- | ···················· | ···················· | ···················· | ···················· | ···················· | ···················· |
| M | 6455 hCoV-19/Scotland/CVR4466/2020\|EPI_ISL_585330\|2020- | ···················· | ···················· | ···················· | ···················· | ······N············· | ··N················· |
| M | 6456 hCoV-19/Scotland/CVR4468/2020\|EPI_ISL_585332\|2020- | ···················· | ···················· | ···················· | ···················· | ···················· | ···················· |
| M | 6457 hCoV-19/Scotland/CVR4469/2020\|EPI_ISL_585333\|2020- | ···················· | ···················· | ···················· | ···················· | ···················· | ···················· |
| M | 6458 hCoV-19/Scotland/CVR4487/2020\|EPI_ISL_585351\|2020- | ···················· | ···················· | ···················· | ···················· | ···················· | ···················· |
| M | 6459 hCoV-19/Scotland/CVR4488/2020\|EPI_ISL_585352\|2020- | ···················· | ···················· | ···················· | ···················· | ···················· | ···················· |
| M | 6460 hCoV-19/Scotland/CVR4490/2020\|EPI_ISL_585353\|2020- | ···················· | ···················· | ···················· | ···················· | ···················· | ···················· |
| M | 6462 hCoV-19/Scotland/CVR4518/2020\|EPI_ISL_585378\|2020- | ···················· | ···················· | ···················· | ···················· | ···················· | ···················· |
| M | 6463 hCoV-19/Scotland/CVR4521/2020\|EPI_ISL_585381\|2020- | ···················· | ···················· | ···················· | ···················· | ···················· | ···················· |
| M | 6464 hCoV-19/Scotland/CVR4525/2020\|EPI_ISL_585385\|2020- | ···················· | ···················· | ···················· | ···················· | ···················· | ···················· |
| M | 6465 hCoV-19/Scotland/CVR4527/2020\|EPI_ISL_585387\|2020- | ···················· | ···················· | ···················· | ···················· | ···················· | ···················· |
| M | 6466 hCoV-19/Scotland/CVR4529/2020\|EPI_ISL_585389\|2020- | ···················· | ···················· | ···················· | ···················· | ···················· | ···················· |
| M | 6467 hCoV-19/Scotland/CVR4531/2020\|EPI_ISL_585391\|2020- | ···················· | ···················· | ···················· | ···················· | ···················· | ···················· |
| M | 6468 hCoV-19/Scotland/CVR4532/2020\|EPI_ISL_585392\|2020- | ···················· | ···················· | ···················· | ···················· | ···················· | ···················· |
| M | 6469 hCoV-19/Scotland/CVR4533/2020\|EPI_ISL_585393\|2020- | ···················· | ···················· | ···················· | ···················· | ···················· | ···················· |
| M | 6470 hCoV-19/Scotland/CVR4534/2020\|EPI_ISL_585394\|2020- | ···················· | ···················· | ···················· | ···················· | ···················· | ···················· |
| M | 6471 hCoV-19/Scotland/CVR4536/2020\|EPI_ISL_585396\|2020- | ···················· | ···················· | ···················· | ···················· | ···················· | ···················· |
| M | 6472 hCoV-19/Scotland/CVR4537/2020\|EPI_ISL_585397\|2020- | ···················· | ···················· | ···················· | ···················· | ···················· | ···················· |
| M | 6473 hCoV-19/England/ALDP-A2E5A9/2020\|EPI_ISL_586600\|20 | ···················· | ···················· | ···················· | ···················· | ···················· | ···················· |
| M | 6474 hCoV-19/England/ALDP-A2DA05/2020\|EPI_ISL_586601\|20 | ···················· | ···················· | ···················· | ···················· | ···················· | ···················· |
| M | 6475 hCoV-19/England/ALDP-A2E011/2020\|EPI_ISL_586602\|20 | ···················· | ···················· | ···················· | ···················· | ···················· | ···················· |
| M | 6476 hCoV-19/England/ALDP-A2DD57/2020\|EPI_ISL_586603\|20 | ···················· | ···················· | ···················· | ···················· | ···················· | ···················· |
| M | 6477 hCoV-19/England/ALDP-A2D5F5/2020\|EPI_ISL_586605\|20 | ···················· | ···················· | ···················· | ···················· | ···················· | ···················· |
| M | 6478 hCoV-19/England/ALDP-A2E0A8/2020\|EPI_ISL_586606\|20 | ···················· | ···················· | ···················· | ···················· | ···················· | ···················· |
| M | 6479 hCoV-19/England/ALDP-A2D179/2020\|EPI_ISL_586607\|20 | ···················· | ···················· | ···················· | ···················· | ···················· | ···················· |
| M | 6480 hCoV-19/England/ALDP-A2DFBB/2020\|EPI_ISL_586608\|20 | ···················· | ···················· | ···················· | ···················· | ···················· | ···················· |
| M | 6481 hCoV-19/England/ALDP-A2D13D/2020\|EPI_ISL_586609\|20 | ···················· | ···················· | ···················· | ···················· | ···················· | ···················· |
| M | 6482 hCoV-19/England/ALDP-A2E2DF/2020\|EPI_ISL_586610\|20 | ···················· | ···················· | ···················· | ···················· | ···················· | ···················· |
| M | 6483 hCoV-19/England/ALDP-A2D9DB/2020\|EPI_ISL_586611\|20 | ···················· | ···················· | ···················· | ···················· | ···················· | ···················· |
| M | 6484 hCoV-19/England/ALDP-A2E49D/2020\|EPI_ISL_586613\|20 | ···················· | ···················· | ···················· | ···················· | ···················· | ···················· |
| M | 6485 hCoV-19/England/ALDP-A2E7C1/2020\|EPI_ISL_586614\|20 | ···················· | ···················· | ···················· | ···················· | ···················· | ···················· |
| M | 6486 hCoV-19/England/ALDP-A2D2FE/2020\|EPI_ISL_586615\|20 | ···················· | ···················· | ···················· | ···················· | ···················· | ···················· |
| M | 6487 hCoV-19/England/ALDP-A2E7D0/2020\|EPI_ISL_586616\|20 | ···················· | ···················· | ···················· | ···················· | ···················· | ···················· |
| M | 6488 hCoV-19/England/ALDP-A2D768/2020\|EPI_ISL_586618\|20 | ···················· | ···················· | ···················· | ···················· | ···················· | ···················· |
| M | 6489 hCoV-19/England/ALDP-A2D72C/2020\|EPI_ISL_586619\|20 | ···················· | ···················· | ···················· | ···················· | ···················· | ···················· |
| M | 6490 hCoV-19/England/ALDP-A2D5AA/2020\|EPI_ISL_586620\|20 | ···················· | ···················· | ···················· | ···················· | ···················· | ···················· |
| M | 6491 hCoV-19/England/ALDP-A2D908/2020\|EPI_ISL_586622\|20 | ···················· | ···················· | ···················· | ···················· | ···················· | ···················· |
| M | 6492 hCoV-19/England/ALDP-A2D55F/2020\|EPI_ISL_586623\|20 | ···················· | ···················· | ···················· | ···················· | ···················· | ···················· |
| M | 6493 hCoV-19/England/ALDP-A2E13C/2020\|EPI_ISL_586624\|20 | ···················· | ···················· | ···················· | ···················· | ···················· | ···················· |
| M | 6494 hCoV-19/England/ALDP-A2D294/2020\|EPI_ISL_586625\|20 | ···················· | ···················· | ···················· | ···················· | ···················· | ···················· |
| M | 6495 hCoV-19/England/ALDP-A2E0C6/2020\|EPI_ISL_586626\|20 | ···················· | ···················· | ···················· | ···················· | ···················· | ···················· |
| M | 6496 hCoV-19/England/ALDP-A2DFF7/2020\|EPI_ISL_586627\|20 | ···················· | ···················· | ···················· | ···················· | ···················· | ···················· |
| M | 6497 hCoV-19/England/ALDP-A2E2C0/2020\|EPI_ISL_586628\|20 | ···················· | ···················· | ·············A······ | ···················· | ···················· | ···················· |
| M | 6498 hCoV-19/England/ALDP-A2DABA/2020\|EPI_ISL_586630\|20 | ···················· | ···················· | ···················· | ···················· | ···················· | ···················· |
| M | 6499 hCoV-19/England/ALDP-A2D9CC/2020\|EPI_ISL_586631\|20 | ···················· | ···················· | ···················· | ···················· | ···················· | ···················· |
| M | 6500 hCoV-19/England/ALDP-A2D971/2020\|EPI_ISL_586632\|20 | ···················· | ·A·················· | ···················· | ···················· | ···················· | ··A················· |
| M | 6501 hCoV-19/England/ALDP-A2DE54/2020\|EPI_ISL_586633\|20 | ···················· | ···················· | ···················· | ···················· | ···················· | ···················· |
| M | 6502 hCoV-19/England/ALDP-A2DE27/2020\|EPI_ISL_586634\|20 | ···················· | ···················· | ···················· | ···················· | ···················· | ···················· |
| M | 6503 hCoV-19/England/ALDP-A2DEFA/2020\|EPI_ISL_586635\|20 | ···················· | ···················· | ···················· | ···················· | ···················· | ···················· |
| M | 6504 hCoV-19/England/ALDP-A2D9BD/2020\|EPI_ISL_586636\|20 | ···················· | ···················· | ···················· | ···················· | ···················· | ···················· |
| M | 6505 hCoV-19/England/ALDP-A2D847/2020\|EPI_ISL_586637\|20 | ···················· | ···················· | ···················· | ···················· | ···················· | ···················· |
| M | 6506 hCoV-19/England/ALDP-A2D1F1/2020\|EPI_ISL_586638\|20 | ···················· | ···················· | ···················· | ···················· | ···················· | ···················· |
| M | 6507 hCoV-19/England/ALDP-A2D276/2020\|EPI_ISL_586639\|20 | ···················· | ···················· | ···················· | ···················· | ···················· | ···················· |
| M | 6508 hCoV-19/England/ALDP-A2E07B/2020\|EPI_ISL_586640\|20 | ···················· | ···················· | ···················· | ···················· | ···················· | ···················· |
| M | 6509 hCoV-19/England/ALDP-A2D267/2020\|EPI_ISL_586641\|20 | ···················· | ···················· | ···················· | ···················· | ···················· | ···················· |
| M | 6510 hCoV-19/England/ALDP-A2D0F4/2020\|EPI_ISL_586642\|20 | ···················· | ···················· | ···················· | ···················· | ···················· | ···················· |
| M | 6511 hCoV-19/England/ALDP-A2DAAB/2020\|EPI_ISL_586643\|20 | ···················· | ···················· | ···················· | ···················· | ···················· | ···················· |
| M | 6512 hCoV-19/England/ALDP-A2DEEB/2020\|EPI_ISL_586644\|20 | ···················· | ···················· | ···················· | ···················· | ···················· | ···················· |
| M | 6513 hCoV-19/England/ALDP-A2E21B/2020\|EPI_ISL_586645\|20 | ···················· | ···················· | ···················· | ···················· | ···················· | ···················· |
| M | 6514 hCoV-19/England/ALDP-A2E7B2/2020\|EPI_ISL_586646\|20 | ···················· | ···················· | ···················· | ···················· | ···················· | ···················· |
| M | 6515 hCoV-19/England/ALDP-A2D80B/2020\|EPI_ISL_586647\|20 | ···················· | ···················· | ···················· | ···················· | ···················· | ···················· |
| M | 6516 hCoV-19/England/ALDP-A2D8A1/2020\|EPI_ISL_586648\|20 | ···················· | ···················· | ···················· | ···················· | ···················· | ···················· |
| M | 6517 hCoV-19/England/ALDP-A2D64D/2020\|EPI_ISL_586649\|20 | ···················· | ···················· | ···················· | ···················· | ···················· | ···················· |
| M | 6518 hCoV-19/England/ALDP-A2E433/2020\|EPI_ISL_586650\|20 | ···················· | ···················· | ···················· | ···················· | ···················· | ···················· |
| M | 6519 hCoV-19/England/ALDP-A2E372/2020\|EPI_ISL_586651\|20 | ···················· | ···················· | ···················· | ···················· | ···················· | ···················· |
| M | 6520 hCoV-19/England/ALDP-A2D74A/2020\|EPI_ISL_586652\|20 | ···················· | ···················· | ···················· | ···················· | ···················· | ···················· |
| M | 6521 hCoV-19/England/ALDP-A2E345/2020\|EPI_ISL_586653\|20 | ···················· | ···················· | ···················· | ···················· | ···················· | ···················· |
| M | 6522 hCoV-19/England/ALDP-A2D73B/2020\|EPI_ISL_586654\|20 | ···················· | ···················· | ···················· | ···················· | ···················· | ···················· |
| M | 6523 hCoV-19/England/ALDP-A2E57C/2020\|EPI_ISL_586655\|20 | ···················· | ···················· | ···················· | ···················· | ···················· | ···················· |
| M | 6524 hCoV-19/England/ALDP-A2E6E2/2020\|EPI_ISL_586656\|20 | ···················· | ···················· | ···················· | ···················· | ···T················ | ···················· |
| M | 6525 hCoV-19/England/ALDP-A2DEDC/2020\|EPI_ISL_586657\|20 | ···················· | ···················· | ···················· | ···················· | ···················· | ···················· |
| M | 6526 hCoV-19/England/ALDP-A2D16A/2020\|EPI_ISL_586659\|20 | ···················· | ···················· | ···················· | ···················· | ···················· | ···················· |
| M | 6527 hCoV-19/Wales/ALDP-A2D531/2020\|EPI_ISL_586660\|2020 | ···················· | ···················· | ···················· | ···················· | ···················· | ···················· |
| M | 6528 hCoV-19/England/ALDP-A2D1A6/2020\|EPI_ISL_586661\|20 | ···················· | ···················· | ···················· | ···················· | ···················· | ···················· |
| M | 6529 hCoV-19/England/ALDP-A2E5D6/2020\|EPI_ISL_586663\|20 | ···················· | ···················· | ···················· | ···················· | ···················· | ···················· |
| M | 6530 hCoV-19/England/ALDP-A2D6D4/2020\|EPI_ISL_586664\|20 | ···················· | ···················· | ···················· | ···················· | ···················· | ···················· |

| | | | | | | | |
|---|---|---|---|---|---|---|---|
| M | 6531 hCoV-19/England/ALDP-A2E06C/2020\|EPI_ISL_586681\|20 | ···················· | ···················· | ···················· | ···················· | ···················· | ···················· |
| M | 6532 hCoV-19/England/ALDP-A2DD66/2020\|EPI_ISL_586682\|20 | ···················· | ···················· | ···················· | ···················· | ···················· | ···················· |
| M | 6533 hCoV-19/England/ALDP-A2DDA2/2020\|EPI_ISL_586683\|20 | ···················· | ···················· | ···················· | ···················· | ···················· | ···················· |
| M | 6534 hCoV-19/England/ALDP-A2DC96/2020\|EPI_ISL_586684\|20 | ···················· | ···················· | ···················· | ···················· | ···················· | ···················· |
| M | 6535 hCoV-19/England/ALDP-A2D382/2020\|EPI_ISL_586685\|20 | ···················· | ···················· | ···················· | ···················· | ···················· | ···················· |
| M | 6536 hCoV-19/England/ALDP-A2E08A/2020\|EPI_ISL_586686\|20 | ···················· | ···················· | ···················· | ···················· | ···················· | ···················· |
| M | 6537 hCoV-19/England/ALDP-9FCAF1/2020\|EPI_ISL_587910\|20 | ···················· | ···················· | ···················· | ···················· | ···················· | ···················· |
| M | 6538 hCoV-19/England/ALDP-9FD4F2/2020\|EPI_ISL_587913\|20 | ···················· | ·A·················· | ···················· | ···················· | ···················· | ···················· |
| M | 6539 hCoV-19/England/ALDP-9FD823/2020\|EPI_ISL_587914\|20 | ···················· | ···················· | ···················· | ···················· | ···················· | ···················· |
| M | 6540 hCoV-19/England/ALDP-9FD96C/2020\|EPI_ISL_587915\|20 | ···················· | ···················· | ···················· | ···················· | ···················· | ···················· |
| M | 6541 hCoV-19/England/ALDP-9FDF2E/2020\|EPI_ISL_587916\|20 | ···················· | ···················· | ···················· | ···················· | ···················· | ···················· |
| M | 6542 hCoV-19/England/ALDP-9FCF01/2020\|EPI_ISL_587917\|20 | ···················· | ···················· | ···················· | ···················· | ···················· | ···················· |
| M | 6543 hCoV-19/England/ALDP-9FDA3C/2020\|EPI_ISL_587918\|20 | ···················· | ···················· | ···················· | ···················· | ···················· | ···················· |
| M | 6544 hCoV-19/England/ALDP-9FD0FE/2020\|EPI_ISL_587919\|20 | ···················· | ···················· | ···················· | ···················· | ······T············· | ···················· |
| M | 6545 hCoV-19/England/ALDP-9FC8E8/2020\|EPI_ISL_587920\|20 | ···················· | ···················· | ···················· | ···················· | ···················· | ···················· |
| M | 6546 hCoV-19/England/ALDP-9FD173/2020\|EPI_ISL_587921\|20 | ···················· | ···················· | ···················· | ···················· | ····T··············· | ···················· |
| M | 6547 hCoV-19/England/ALDP-9FDE5E/2020\|EPI_ISL_587922\|20 | ···················· | ···················· | ···················· | ···················· | ···················· | ···················· |
| M | 6548 hCoV-19/England/ALDP-9FCAE2/2020\|EPI_ISL_587923\|20 | ···················· | ···················· | ···················· | ···················· | ···················· | ···················· |
| M | 6549 hCoV-19/England/ALDP-9FD6ED/2020\|EPI_ISL_587924\|20 | ···················· | ···················· | ···················· | ···················· | ···················· | ···················· |
| M | 6550 hCoV-19/England/ALDP-9FD9D5/2020\|EPI_ISL_587926\|20 | ···················· | ·A·················· | ···················· | ···················· | ···················· | ···A················ |
| M | 6551 hCoV-19/England/ALDP-9FCA4C/2020\|EPI_ISL_587927\|20 | ···················· | ···················· | ···················· | ···················· | ···················· | ···················· |
| M | 6552 hCoV-19/England/ALDP-9FD52C/2020\|EPI_ISL_587930\|20 | ···················· | ···················· | ···················· | ···················· | ···················· | ···················· |
| M | 6553 hCoV-19/England/ALDP-9FDD60/2020\|EPI_ISL_587931\|20 | ···················· | ···················· | ···················· | ···················· | ···················· | ···················· |
| M | 6554 hCoV-19/England/ALDP-9FCC82/2020\|EPI_ISL_587932\|20 | ···················· | ···················· | ···················· | ···················· | ···················· | ···················· |
| M | 6555 hCoV-19/England/ALDP-9FCBC1/2020\|EPI_ISL_587933\|20 | ···················· | ···················· | ···················· | ···················· | ···················· | ···················· |
| M | 6556 hCoV-19/England/ALDP-9FCC64/2020\|EPI_ISL_587934\|20 | ···················· | ···················· | ···················· | ···················· | ···················· | ···················· |
| M | 6557 hCoV-19/England/ALDP-9FC903/2020\|EPI_ISL_587935\|20 | ···················· | ···················· | ···················· | ···················· | ···················· | ···················· |
| M | 6558 hCoV-19/England/ALDP-9FD085/2020\|EPI_ISL_587936\|20 | ···················· | ···················· | ···················· | ···················· | ···················· | ···················· |
| M | 6559 hCoV-19/England/ALDP-9FDB0C/2020\|EPI_ISL_587937\|20 | ···················· | ···················· | ···················· | ···················· | ···················· | ···················· |
| M | 6560 hCoV-19/England/ALDP-9FD665/2020\|EPI_ISL_587938\|20 | ···················· | ···················· | ···················· | ···················· | ···················· | ···················· |
| M | 6561 hCoV-19/England/ALDP-9FD7F9/2020\|EPI_ISL_587939\|20 | ···················· | ···················· | ···················· | ···················· | ···················· | ···················· |
| M | 6562 hCoV-19/England/ALDP-9FD902/2020\|EPI_ISL_587940\|20 | ···················· | ···················· | ···················· | ···················· | ···················· | ···················· |
| M | 6563 hCoV-19/England/ALDP-9FDE9A/2020\|EPI_ISL_587941\|20 | ···················· | ···················· | ···················· | ···················· | ···················· | ···················· |
| M | 6564 hCoV-19/England/ALDP-9FD39B/2020\|EPI_ISL_587942\|20 | ···················· | ···················· | ···················· | ···················· | ···················· | ···················· |
| M | 6565 hCoV-19/England/ALDP-9FDC27/2020\|EPI_ISL_587943\|20 | ···················· | ···················· | ···················· | ···················· | ···················· | ···················· |
| M | 6566 hCoV-19/England/ALDP-9FCA2E/2020\|EPI_ISL_587944\|20 | ···················· | ·A·················· | ···················· | ···················· | ···················· | ···A················ |
| M | 6567 hCoV-19/England/ALDP-9FDC45/2020\|EPI_ISL_587945\|20 | ···················· | ···················· | ···················· | ···················· | ···················· | ···················· |
| M | 6568 hCoV-19/England/ALDP-9FDF88/2020\|EPI_ISL_587946\|20 | ···················· | ···················· | ···················· | ···················· | ···················· | ···················· |
| M | 6569 hCoV-19/England/ALDP-9FDC09/2020\|EPI_ISL_587948\|20 | ···················· | ···················· | ···················· | ···················· | ···················· | ···················· |
| M | 6570 hCoV-19/England/ALDP-9FD10A/2020\|EPI_ISL_587949\|20 | ···················· | ···················· | ···················· | ···················· | ···················· | ···················· |
| M | 6571 hCoV-19/England/ALDP-9FD4E3/2020\|EPI_ISL_587951\|20 | ···················· | ···················· | ···················· | ···················· | ···················· | ···················· |
| M | 6572 hCoV-19/England/ALDP-9FD5C2/2020\|EPI_ISL_587953\|20 | ···················· | ···················· | ···················· | ···················· | ···················· | ···················· |
| M | 6573 hCoV-19/England/ALDP-9FDBEE/2020\|EPI_ISL_587954\|20 | ···················· | ···················· | ···················· | ···················· | ····T··············· | ···················· |
| M | 6574 hCoV-19/England/ALDP-9FD8AB/2020\|EPI_ISL_587955\|20 | ···················· | ···················· | ···················· | ···················· | ···················· | ···················· |
| M | 6575 hCoV-19/England/ALDP-9FD29E/2020\|EPI_ISL_587956\|20 | ···················· | ·A·················· | ···················· | ···················· | ···················· | ···················· |
| M | 6576 hCoV-19/England/ALDP-9FD692/2020\|EPI_ISL_587957\|20 | ···················· | ···················· | ···················· | ···················· | ···················· | ···················· |
| M | 6577 hCoV-19/England/ALDP-9FC851/2020\|EPI_ISL_587958\|20 | ···················· | ···················· | ···················· | ···················· | ···················· | ···················· |
| M | 6578 hCoV-19/England/ALDP-9FDBB1/2020\|EPI_ISL_587961\|20 | ···················· | ···················· | ···················· | ···················· | ···················· | ···················· |
| M | 6579 hCoV-19/England/ALDP-9FD03A/2020\|EPI_ISL_587963\|20 | ···················· | ···················· | ···················· | ···················· | ···················· | ···················· |
| M | 6580 hCoV-19/England/ALDP-9FD36E/2020\|EPI_ISL_587964\|20 | ···················· | ···················· | ···················· | ···················· | ···················· | ···················· |
| M | 6581 hCoV-19/England/ALDP-9FD5E0/2020\|EPI_ISL_587965\|20 | ···················· | ···················· | ···················· | ···················· | ···················· | ···················· |
| M | 6582 hCoV-19/England/ALDP-9FCE9B/2020\|EPI_ISL_587966\|20 | ···················· | ···················· | ···················· | ···················· | ···················· | ···················· |
| M | 6583 hCoV-19/England/ALDP-9FCFB6/2020\|EPI_ISL_587967\|20 | ···················· | ···················· | ···················· | ···················· | ···················· | ···················· |
| M | 6584 hCoV-19/England/ALDP-9FD42F/2020\|EPI_ISL_587968\|20 | ···················· | ···················· | ···················· | ···················· | ···················· | ···················· |
| M | 6585 hCoV-19/England/ALDP-9FD270/2020\|EPI_ISL_587969\|20 | ···················· | ···················· | ···················· | ···················· | ···················· | ···················· |
| M | 6586 hCoV-19/England/ALDP-9FD999/2020\|EPI_ISL_587970\|20 | ···················· | ···················· | ···················· | ···················· | ···················· | ···················· |
| M | 6587 hCoV-19/England/ALDP-9FD4A7/2020\|EPI_ISL_587971\|20 | ···················· | ···················· | ···················· | ···················· | ···················· | ···················· |
| M | 6588 hCoV-19/England/ALDP-9FDC63/2020\|EPI_ISL_587975\|20 | ···················· | ···················· | ···················· | ···················· | ···················· | ···················· |
| M | 6589 hCoV-19/England/ALDP-9FD726/2020\|EPI_ISL_587976\|20 | ···················· | ···················· | ···················· | ···················· | ···················· | ···················· |
| M | 6590 hCoV-19/England/ALDP-9FE01B/2020\|EPI_ISL_587978\|20 | ···················· | ···················· | ···················· | ···················· | ···················· | ···················· |
| M | 6591 hCoV-19/England/ALDP-9FD95D/2020\|EPI_ISL_587979\|20 | ···················· | ···················· | ···················· | ···················· | ···················· | ···················· |
| M | 6592 hCoV-19/England/ALDP-9FD094/2020\|EPI_ISL_587980\|20 | ···················· | ···················· | ···················· | ···················· | ···················· | ···················· |
| M | 6593 hCoV-19/England/ALDP-9FCB76/2020\|EPI_ISL_587981\|20 | ···················· | ···················· | ···················· | ···················· | ···················· | ···················· |
| M | 6594 hCoV-19/England/ALDP-9FD2F8/2020\|EPI_ISL_587982\|20 | ···················· | ···················· | ···················· | ···················· | ···················· | ···················· |
| M | 6595 hCoV-19/England/ALDP-9FD9A8/2020\|EPI_ISL_587983\|20 | ···················· | ···················· | ···················· | ···················· | ······T············· | ···················· |
| M | 6596 hCoV-19/England/ALDP-9FCBA3/2020\|EPI_ISL_587984\|20 | ···················· | ···················· | ···················· | ···················· | ···················· | ···················· |
| M | 6597 hCoV-19/England/ALDP-9FD54A/2020\|EPI_ISL_587986\|20 | ···················· | ···················· | ···················· | ···················· | ···················· | ···················· |
| M | 6598 hCoV-19/England/ALDP-9FC824/2020\|EPI_ISL_587987\|20 | ···················· | ···················· | ···················· | ···················· | ···················· | ···················· |
| M | 6599 hCoV-19/England/ALDP-9FD2E9/2020\|EPI_ISL_587988\|20 | ···················· | ·R·················· | ···················· | ···················· | ···················· | ···R················ |
| M | 6600 hCoV-19/England/ALDP-9FD3AA/2020\|EPI_ISL_587989\|20 | ···················· | ···················· | ···················· | ···················· | ···················· | ···················· |
| M | 6601 hCoV-19/England/ALDP-9FCF2F/2020\|EPI_ISL_587990\|20 | ···················· | ···················· | ···················· | ···················· | ···················· | ···················· |
| M | 6602 hCoV-19/England/ALDP-9FD9F3/2020\|EPI_ISL_587991\|20 | ···················· | ···················· | ···················· | ···················· | ···················· | ···················· |
| M | 6603 hCoV-19/England/ALDP-9FCAB5/2020\|EPI_ISL_587992\|20 | ···················· | ···················· | ···················· | ···················· | ···················· | ···················· |
| M | 6604 hCoV-19/England/ALDP-9FCE22/2020\|EPI_ISL_587993\|20 | ···················· | ···················· | ···················· | ···················· | ···················· | ···················· |
| M | 6605 hCoV-19/England/ALDP-9FCCFB/2020\|EPI_ISL_587994\|20 | ···················· | ···················· | ···················· | ···················· | ···················· | ···················· |
| M | 6606 hCoV-19/England/ALDP-9FD93F/2020\|EPI_ISL_587995\|20 | ···················· | ···················· | ···················· | ···················· | ···················· | ···················· |
| M | 6607 hCoV-19/Canada/ON-S878/2020\|EPI_ISL_591175\|2020-04 | ···················· | ···················· | ···················· | ···················· | ···················· | ···················· |
| M | 6608 hCoV-19/Canada/ON-S879/2020\|EPI_ISL_591176\|2020-04 | ···················· | ···················· | ···················· | ···················· | ···················· | ···················· |
| M | 6609 hCoV-19/Canada/ON-S895/2020\|EPI_ISL_591177\|2020-04 | ···················· | ···················· | ···················· | ···················· | ···················· | ···················· |
| M | 6610 hCoV-19/Canada/ON-S896/2020\|EPI_ISL_591178\|2020-04 | ···················· | ···················· | ···················· | ···················· | ···················· | ···················· |

| | | | | | | | | |
|---|---|---|---|---|---|---|---|---|
| M | 6611 hCoV-19/Canada/ON-S921/2020\|EPI_ISL_591179\|2020-04 | ···················· | ···················· | ···················· | ···················· | ···················· | ···················· | |
| M | 6612 hCoV-19/Canada/ON-S957/2020\|EPI_ISL_591181\|2020-04 | ···················· | ···················· | ···················· | ···················· | ···················· | ···················· | |
| M | 6613 hCoV-19/Canada/ON-S125/2020\|EPI_ISL_591182\|2020-03 | ···················· | ···················· | ···················· | ···················· | ···················· | ···················· | |
| M | 6614 hCoV-19/Canada/ON-S1415/2020\|EPI_ISL_591186\|2020-0 | ···················· | ···················· | ···················· | ···················· | ···················· | ···················· | |
| M | 6615 hCoV-19/Canada/ON-S1416/2020\|EPI_ISL_591187\|2020-0 | ···················· | ···················· | ···················· | ···················· | ···················· | ···················· | |
| M | 6616 hCoV-19/Canada/ON-S1427/2020\|EPI_ISL_591193\|2020-0 | ···················· | ···················· | ···················· | ···················· | ···················· | ···················· | |
| M | 6617 hCoV-19/Canada/ON-S1434/2020\|EPI_ISL_591197\|2020-0 | ···················· | ···················· | ···················· | ···················· | ···················· | ···················· | |
| M | 6618 hCoV-19/Canada/ON-S1436/2020\|EPI_ISL_591199\|2020-0 | ···················· | ···················· | ···················· | ···················· | ···················· | ···················· | |
| M | 6619 hCoV-19/Australia/VIC13812/2020\|EPI_ISL_592325\|202 | ···················· | ···················· | ···················· | ···················· | ···················· | ···················· | |
| M | 6620 hCoV-19/Australia/VIC13815/2020\|EPI_ISL_592326\|202 | ···················· | ···················· | ···················· | ···················· | ···················· | ···················· | |
| M | 6621 hCoV-19/Australia/NSW1079/2020\|EPI_ISL_593654\|2020 | ···················· | ···················· | ···················· | ···················· | ···················· | ···················· | |
| M | 6622 hCoV-19/Australia/NSW1070/2020\|EPI_ISL_593656\|2020 | ···················· | ···················· | ···················· | ···················· | ···················· | ···················· | |
| M | 6623 hCoV-19/Australia/NSW1072/2020\|EPI_ISL_593657\|2020 | ···················· | ···················· | ···················· | ···················· | ···················· | ···················· | |
| M | 6624 hCoV-19/Australia/NSW1015/2020\|EPI_ISL_593660\|2020 | ···················· | ···················· | ···················· | ···················· | ···················· | ···················· | |
| M | 6625 hCoV-19/England/MILK-A17DAB/2020\|EPI_ISL_594987\|20 | ···················· | ···················· | ···················· | ···················· | ···················· | ···················· | |
| M | 6626 hCoV-19/England/MILK-A17DC9/2020\|EPI_ISL_594988\|20 | ···················· | ···················· | ···················· | ···················· | ···················· | ···················· | |
| M | 6627 hCoV-19/England/MILK-A17E02/2020\|EPI_ISL_594990\|20 | ···················· | ···················· | ···················· | ···················· | ···················· | ···················· | |
| M | 6628 hCoV-19/England/MILK-A17E20/2020\|EPI_ISL_594991\|20 | ···················· | ···················· | ···················· | ···················· | ···················· | ···················· | |
| M | 6629 hCoV-19/England/QEUH-A4CB76/2020\|EPI_ISL_596026\|20 | ···················· | ···················· | ···················· | ···················· | ···················· | ···················· | |
| M | 6630 hCoV-19/England/QEUH-A4CB85/2020\|EPI_ISL_596027\|20 | ···················· | ·A·················· | ···················· | ···················· | ···················· | ··A················· | |
| M | 6631 hCoV-19/England/QEUH-A4CBEF/2020\|EPI_ISL_596031\|20 | ···················· | ···················· | ···················· | ···················· | ···················· | ···················· | |
| M | 6632 hCoV-19/Scotland/QEUH-A4CC0A/2020\|EPI_ISL_596033\|2 | ···················· | ···················· | ···················· | ···················· | ···················· | ···················· | |
| M | 6633 hCoV-19/Scotland/QEUH-A4CC19/2020\|EPI_ISL_596034\|2 | ···················· | ···················· | ···················· | ···················· | ···················· | ···················· | |
| M | 6635 hCoV-19/Scotland/QEUH-A4CC46/2020\|EPI_ISL_596036\|2 | ···················· | ···················· | ···················· | ···················· | ···················· | ···················· | |
| M | 6636 hCoV-19/Scotland/QEUH-A4CC73/2020\|EPI_ISL_596038\|2 | ···················· | ···················· | ···················· | ···················· | ···················· | ···················· | |
| M | 6637 hCoV-19/Scotland/QEUH-A4CC82/2020\|EPI_ISL_596039\|2 | ···················· | ···················· | ···················· | ···················· | ···················· | ···················· | |
| M | 6638 hCoV-19/Scotland/QEUH-A4CCBF/2020\|EPI_ISL_596041\|2 | ···················· | ···················· | ···················· | ···················· | NNN················· | ···················· | 1 |
| M | 6639 hCoV-19/Scotland/QEUH-A4CCFB/2020\|EPI_ISL_596045\|2 | ···················· | ···················· | ···················· | ···················· | ···················· | ···················· | |
| M | 6640 hCoV-19/Scotland/QEUH-A4CD07/2020\|EPI_ISL_596046\|2 | ···················· | ···················· | ···················· | ···················· | ···················· | ···················· | |
| M | 6641 hCoV-19/Scotland/QEUH-A4CD16/2020\|EPI_ISL_596047\|2 | ···················· | ···················· | ···················· | ···················· | ···················· | ···················· | |
| M | 6643 hCoV-19/England/QEUH-A4CD34/2020\|EPI_ISL_596049\|20 | ···················· | ···················· | ···················· | ···················· | ···················· | ···················· | |
| M | 6644 hCoV-19/England/QEUH-A4CD43/2020\|EPI_ISL_596050\|20 | ···················· | ···················· | ···················· | ···················· | NNN················· | ···················· | 1 |
| M | 6645 hCoV-19/England/QEUH-A4CD61/2020\|EPI_ISL_596051\|20 | ···················· | ···················· | ···················· | ···················· | ···················· | ···················· | |
| M | 6646 hCoV-19/England/QEUH-A4CD8F/2020\|EPI_ISL_596053\|20 | ···················· | ···················· | ···················· | ···················· | ···················· | ···················· | |
| M | 6647 hCoV-19/England/QEUH-A4CDBC/2020\|EPI_ISL_596056\|20 | ···················· | ···················· | ···················· | ···················· | ···················· | ···················· | |
| M | 6650 hCoV-19/England/QEUH-A4CE13/2020\|EPI_ISL_596059\|20 | ···················· | ·A·················· | ···················· | ···················· | ···················· | ··A················· | |
| M | 6651 hCoV-19/Scotland/QEUH-A4CEC8/2020\|EPI_ISL_596063\|2 | ···················· | ···················· | ···················· | ···················· | ···················· | ···················· | |
| M | 6652 hCoV-19/England/QEUH-A4CED7/2020\|EPI_ISL_596064\|20 | ···················· | ···················· | ···················· | ···················· | ···················· | ···················· | |
| M | 6653 hCoV-19/England/QEUH-A4CEE6/2020\|EPI_ISL_596065\|20 | ···················· | ···················· | ···················· | ···················· | ···················· | ···················· | |
| M | 6654 hCoV-19/England/QEUH-A4CF01/2020\|EPI_ISL_596067\|20 | ···················· | ·A·················· | ···················· | ···················· | ···················· | ··A················· | |
| M | 6655 hCoV-19/England/QEUH-A4CF2F/2020\|EPI_ISL_596068\|20 | ···················· | ···················· | ···················· | ···················· | ···················· | ···················· | |
| M | 6656 hCoV-19/England/QEUH-A4CF3E/2020\|EPI_ISL_596069\|20 | ···················· | ·A·················· | ···················· | ···················· | ···················· | ··A················· | |
| M | 6660 hCoV-19/Northern Ireland/NIRE-1082D4/2020\|EPI_ISL_ | ···················· | ···················· | ···················· | ···················· | ···················· | ···················· | |
| M | 6661 hCoV-19/Northern Ireland/NIRE-1082F2/2020\|EPI_ISL_ | ···················· | ···················· | ···················· | ···················· | ···················· | ···················· | |
| M | 6662 hCoV-19/Northern Ireland/NIRE-108377/2020\|EPI_ISL_ | ···················· | ···················· | ···················· | ···················· | ···················· | ···················· | |
| M | 6663 hCoV-19/Northern Ireland/NIRE-108395/2020\|EPI_ISL_ | ···················· | ···················· | ···················· | ···················· | ···················· | ···················· | |
| M | 6664 hCoV-19/Northern Ireland/NIRE-1083B3/2020\|EPI_ISL_ | ···················· | ···················· | ···················· | ···················· | ···················· | ···················· | |
| M | 6665 hCoV-19/Northern Ireland/NIRE-1083D1/2020\|EPI_ISL_ | ···················· | ···················· | ···················· | ···················· | ···················· | ···················· | |
| M | 6666 hCoV-19/Northern Ireland/NIRE-1083FF/2020\|EPI_ISL_ | ···················· | ···················· | ···················· | ···················· | ···················· | ···················· | |
| M | 6667 hCoV-19/Northern Ireland/NIRE-108438/2020\|EPI_ISL_ | ···················· | ···················· | ···················· | ···················· | ···················· | ···················· | |
| M | 6668 hCoV-19/Northern Ireland/NIRE-108456/2020\|EPI_ISL_ | ···················· | ···················· | ···················· | ···················· | ···················· | ···················· | |
| M | 6669 hCoV-19/Northern Ireland/NIRE-108614/2020\|EPI_ISL_ | ···················· | ···················· | ···················· | ···················· | ···················· | ···················· | |
| M | 6670 hCoV-19/Northern Ireland/NIRE-108650/2020\|EPI_ISL_ | ···················· | ···················· | ···················· | ···················· | ···················· | ···················· | |
| M | 6671 hCoV-19/Northern Ireland/NIRE-1086F6/2020\|EPI_ISL_ | ···················· | ···················· | ···················· | ···················· | ···················· | ···················· | |
| M | 6672 hCoV-19/Northern Ireland/NIRE-108711/2020\|EPI_ISL_ | ···················· | ···················· | ···················· | ···················· | ···················· | ···················· | |
| M | 6673 hCoV-19/Northern Ireland/NIRE-10875D/2020\|EPI_ISL_ | ···················· | ···················· | ···················· | ···················· | ···················· | ···················· | |
| M | 6674 hCoV-19/Norway/3745/2020\|EPI_ISL_591000\|2020-09-04 | ···················· | ···················· | ···················· | ···················· | ···················· | ···················· | |
| M | 6675 hCoV-19/Norway/3747/2020\|EPI_ISL_591001\|2020-09-03 | ···················· | ···················· | ···················· | ···················· | ···················· | ···················· | |
| M | 6676 hCoV-19/Norway/3750/2020\|EPI_ISL_591002\|2020-09-05 | ···················· | ···················· | ···················· | ···················· | ···················· | ···················· | |
| M | 6677 hCoV-19/Norway/3753/2020\|EPI_ISL_591003\|2020-09-08 | ···················· | ···················· | ···················· | ···················· | ···················· | ···················· | |
| M | 6678 hCoV-19/Norway/3754/2020\|EPI_ISL_591004\|2020-09-06 | ···················· | ···················· | ···················· | ···················· | ···················· | ···················· | |
| M | 6679 hCoV-19/Norway/3755/2020\|EPI_ISL_591005\|2020-09-07 | ···················· | ···················· | ···················· | ···················· | ···················· | ···················· | |
| M | 6680 hCoV-19/Norway/3756/2020\|EPI_ISL_591006\|2020-08-14 | ···················· | ···················· | ···················· | ···················· | ···················· | ···················· | |
| M | 6681 hCoV-19/Norway/3629/2020\|EPI_ISL_591007\|2020-08-06 | ···················· | ·A·················· | ···················· | ···················· | ···················· | ··A················· | |
| M | 6682 hCoV-19/Norway/3841/2020\|EPI_ISL_591008\|2020-09-28 | ···················· | ···················· | ···················· | ···················· | ···················· | ···················· | |
| M | 6683 hCoV-19/Norway/3844/2020\|EPI_ISL_591009\|2020-09-28 | ···················· | ···················· | ···················· | ···················· | ···················· | ···················· | |
| M | 6684 hCoV-19/Norway/3828/2020\|EPI_ISL_591010\|2020-09-24 | ···················· | ···················· | ···················· | ···················· | ···················· | ···················· | |
| M | 6685 hCoV-19/Norway/3833/2020\|EPI_ISL_591011\|2020-09-27 | ···················· | ···················· | ···················· | ···················· | ···················· | ···················· | |
| M | 6686 hCoV-19/Norway/3839/2020\|EPI_ISL_591012\|2020-09-26 | ···················· | ····A··············· | ···················· | ···················· | ···················· | ···················· | |
| M | 6687 hCoV-19/Norway/3829/2020\|EPI_ISL_591013\|2020-09-22 | ···················· | ··········A········· | ···················· | ···················· | ···················· | ···················· | |
| M | 6688 hCoV-19/Norway/3897/2020\|EPI_ISL_591014\|2020-09-30 | ···················· | ···················· | ···················· | ···················· | ···················· | ···················· | |
| M | 6689 hCoV-19/Norway/3722/2020\|EPI_ISL_591015\|2020-09-01 | ···················· | ···················· | ···················· | ···················· | ···················· | ···················· | |
| M | 6690 hCoV-19/Norway/3765/2020\|EPI_ISL_591016\|2020-08-29 | ···················· | ···················· | ···················· | ···················· | ···················· | ···················· | |
| M | 6691 hCoV-19/Norway/3766/2020\|EPI_ISL_591017\|2020-08-29 | ···················· | ···················· | ···················· | ···················· | ···················· | ···················· | |
| M | 6692 hCoV-19/Norway/3913/2020\|EPI_ISL_591018\|2020-09-28 | ···················· | ···················· | ···················· | ···················· | ···················· | ···················· | |
| M | 6693 hCoV-19/Norway/3915/2020\|EPI_ISL_591019\|2020-09-29 | ···················· | ···················· | ···················· | ···················· | ···················· | ···················· | |
| M | 6694 hCoV-19/Norway/3292/2020\|EPI_ISL_591020\|2020-09-03 | ···················· | ···················· | ···················· | ···················· | ···················· | ···················· | |
| M | 6695 hCoV-19/Norway/3806/2020\|EPI_ISL_591021\|2020-08-01 | ···················· | ·A·················· | ···················· | ···················· | ···················· | ··A················· | |
| M | 6696 hCoV-19/USA/MD-MDH-0220/2020\|EPI_ISL_591022\|2020-0 | ···················· | ···················· | ···················· | ···················· | ···················· | ···················· | |
| M | 6697 hCoV-19/USA/MD-MDH-0221/2020\|EPI_ISL_591023\|2020-0 | ···················· | ···················· | ···················· | ···················· | ···················· | ···················· | |

| | | | | | | | | |
|---|---|---|---|---|---|---|---|---|
| M | 6698 | hCoV-19/USA/MD-MDH-0222/2020\|EPI_ISL_591024\|2020-0 | ···················· | ···················· | ···················· | ···················· | ···················· | ···················· |
| M | 6699 | hCoV-19/USA/MD-MDH-0223/2020\|EPI_ISL_591025\|2020-0 | ···················· | ···················· | ···················· | ···················· | ···················· | ···················· |
| M | 6700 | hCoV-19/USA/MD-MDH-0224/2020\|EPI_ISL_591026\|2020-0 | ···················· | ···················· | ···················· | ···················· | ···················· | ···················· |
| M | 6701 | hCoV-19/USA/MD-MDH-0225/2020\|EPI_ISL_591027\|2020-0 | ···················· | ···················· | ···················· | ···················· | ···················· | ···················· |
| M | 6702 | hCoV-19/USA/MD-MDH-0226/2020\|EPI_ISL_591028\|2020-0 | ···················· | ···················· | ···················· | ···················· | ···················· | ···················· |
| M | 6703 | hCoV-19/USA/MD-MDH-0227/2020\|EPI_ISL_591029\|2020-0 | ···················· | ·A·················· | ···················· | ···················· | ···················· | ···················· |
| M | 6704 | hCoV-19/USA/MD-MDH-0228/2020\|EPI_ISL_591030\|2020-0 | ···················· | ···················· | ···················· | ···················· | ···················· | ···················· |
| M | 6705 | hCoV-19/USA/MD-MDH-0229/2020\|EPI_ISL_591031\|2020-0 | ···················· | ···················· | ···················· | ···················· | ···················· | ···················· |
| M | 6706 | hCoV-19/USA/MD-MDH-0230/2020\|EPI_ISL_591032\|2020-0 | ···················· | ···················· | ···················· | ···················· | ···················· | ···················· |
| M | 6707 | hCoV-19/USA/MD-MDH-0232/2020\|EPI_ISL_591034\|2020-0 | ···················· | ···················· | ···················· | ···················· | ···················· | ···················· |
| M | 6708 | hCoV-19/USA/MD-MDH-0233/2020\|EPI_ISL_591035\|2020-0 | ···················· | ·A·················· | ···················· | ···················· | ···················· | ···················· |
| M | 6709 | hCoV-19/USA/MD-MDH-0234/2020\|EPI_ISL_591036\|2020-0 | ···················· | ···················· | ···················· | ···················· | ···················· | ···················· |
| M | 6710 | hCoV-19/Australia/VIC13691/2020\|EPI_ISL_592300\|202 | ···················· | ···················· | ···················· | ···················· | ···················· | ···················· |
| M | 6711 | hCoV-19/Australia/VIC13692/2020\|EPI_ISL_592301\|202 | ···················· | ···················· | ···················· | ···················· | ···················· | ···················· |
| M | 6712 | hCoV-19/Australia/VIC13693/2020\|EPI_ISL_592302\|202 | ···················· | ···················· | ···················· | ···················· | ···················· | ···················· |
| M | 6713 | hCoV-19/Australia/VIC13694/2020\|EPI_ISL_592303\|202 | ···················· | ···················· | ···················· | ···················· | ···················· | ···················· |
| M | 6714 | hCoV-19/Australia/VIC13695/2020\|EPI_ISL_592304\|202 | ···················· | ···················· | ···················· | ···················· | ···················· | ···················· |
| M | 6716 | hCoV-19/Australia/VIC13698/2020\|EPI_ISL_592306\|202 | ···················· | ···················· | ···················· | ···················· | ···················· | ···················· |
| M | 6720 | hCoV-19/Australia/VIC13724/2020\|EPI_ISL_592310\|202 | ···················· | ···················· | ···················· | ···················· | ···················· | ···················· |
| M | 6721 | hCoV-19/Australia/VIC13735/2020\|EPI_ISL_592311\|202 | ···················· | ···················· | ···················· | ···················· | ···················· | ···················· |
| M | 6722 | hCoV-19/Australia/VIC13739/2020\|EPI_ISL_592312\|202 | ···················· | ···················· | ···················· | ···················· | ···················· | ···················· |
| M | 6723 | hCoV-19/Australia/VIC13740/2020\|EPI_ISL_592313\|202 | ···················· | ···················· | ···················· | ···················· | ···················· | ···················· |
| M | 6724 | hCoV-19/Australia/VIC13746/2020\|EPI_ISL_592314\|202 | ···················· | ···················· | ···················· | ···················· | ···················· | ···················· |
| M | 6725 | hCoV-19/Australia/VIC13794/2020\|EPI_ISL_592315\|202 | ···················· | ···················· | ···················· | ···················· | ···················· | ···················· |
| M | 6726 | hCoV-19/Australia/VIC13796/2020\|EPI_ISL_592316\|202 | ···················· | ···················· | ···················· | ···················· | ···················· | ···················· |
| M | 6727 | hCoV-19/Australia/VIC13797/2020\|EPI_ISL_592317\|202 | ···················· | ···················· | ···················· | ···················· | ···················· | ···················· |
| M | 6728 | hCoV-19/Australia/VIC13798/2020\|EPI_ISL_592318\|202 | ···················· | ···················· | ···················· | ···················· | ···················· | ···················· |
| M | 6729 | hCoV-19/Australia/VIC13802/2020\|EPI_ISL_592319\|202 | ···················· | ···················· | ···················· | ···················· | NN·················· | ···················· |
| M | 6730 | hCoV-19/Australia/VIC13804/2020\|EPI_ISL_592320\|202 | ···················· | ···················· | ···················· | ···················· | ···················· | ···················· |
| M | 6731 | hCoV-19/Australia/VIC13805/2020\|EPI_ISL_592321\|202 | ···················· | ···················· | ···················· | ···················· | ···················· | ···················· |
| M | 6732 | hCoV-19/Australia/VIC13806/2020\|EPI_ISL_592322\|202 | ···················· | ···················· | ···················· | ···················· | ···················· | ···················· |
| M | 6733 | hCoV-19/Australia/VIC13808/2020\|EPI_ISL_592323\|202 | ···················· | ···················· | ···················· | ···················· | ···················· | ···················· |
| M | 6734 | hCoV-19/Australia/VIC13809/2020\|EPI_ISL_592324\|202 | ···················· | ···················· | ···················· | ···················· | ···················· | ···················· |
| M | 6735 | hCoV-19/Australia/VIC13818/2020\|EPI_ISL_592327\|202 | ···················· | ···················· | ···················· | ···················· | ···················· | ···················· |
| M | 6736 | hCoV-19/Australia/VIC13832/2020\|EPI_ISL_592328\|202 | ···················· | ···················· | ···················· | ···················· | ···················· | ···················· |
| M | 6738 | hCoV-19/Australia/VIC13840/2020\|EPI_ISL_592330\|202 | ···················· | ···················· | ···················· | ···················· | ···················· | ···················· |
| M | 6739 | hCoV-19/Australia/VIC13842/2020\|EPI_ISL_592331\|202 | ···················· | ···················· | ···················· | ···················· | ···················· | ···················· |
| M | 6740 | hCoV-19/Australia/VIC13847/2020\|EPI_ISL_592332\|202 | ···················· | ···················· | ···················· | ···················· | ···················· | ···················· |
| M | 6742 | hCoV-19/Australia/VIC13865/2020\|EPI_ISL_592334\|202 | ···················· | ···················· | ···················· | ···················· | ···················· | ···················· |
| M | 6743 | hCoV-19/Australia/VIC13872/2020\|EPI_ISL_592335\|202 | ···················· | ···················· | ···················· | ···················· | ···················· | ···················· |
| M | 6745 | hCoV-19/Australia/VIC13875/2020\|EPI_ISL_592337\|202 | ···················· | ···················· | ···················· | ···················· | ···················· | ···················· |
| M | 6746 | hCoV-19/Australia/VIC13876/2020\|EPI_ISL_592338\|202 | ···················· | ···················· | ···················· | ···················· | ···················· | ···················· |
| M | 6747 | hCoV-19/Australia/VIC13884/2020\|EPI_ISL_592339\|202 | ···················· | ···················· | ···················· | ···················· | ···················· | ···················· |
| M | 6748 | hCoV-19/Australia/VIC13887/2020\|EPI_ISL_592340\|202 | ···················· | ···················· | ···················· | ···················· | ···················· | ···················· |
| M | 6753 | hCoV-19/Australia/VIC14078/2020\|EPI_ISL_592345\|202 | ···················· | ···················· | ···················· | ···················· | ···················· | ···················· |
| M | 6754 | hCoV-19/Australia/VIC14118/2020\|EPI_ISL_592346\|202 | ···················· | ···················· | ···················· | ···················· | ···················· | ···················· |
| M | 6757 | hCoV-19/Australia/VIC14130/2020\|EPI_ISL_592349\|202 | ···················· | ···················· | ···················· | ···················· | ···················· | ···················· |
| M | 6758 | hCoV-19/Australia/VIC14132/2020\|EPI_ISL_592350\|202 | ···················· | ···················· | ···················· | ···················· | ···················· | ···················· |
| M | 6759 | hCoV-19/Australia/VIC14138/2020\|EPI_ISL_592351\|202 | ···················· | ···················· | ···················· | ···················· | N··················· | ···················· |
| M | 6760 | hCoV-19/Australia/VIC14149/2020\|EPI_ISL_592352\|202 | ···················· | ···················· | ···················· | ···················· | ···················· | ···················· |
| M | 6761 | hCoV-19/Australia/VIC14150/2020\|EPI_ISL_592353\|202 | ···················· | ···················· | ···················· | ···················· | ···················· | ···················· |
| M | 6762 | hCoV-19/Australia/VIC14151/2020\|EPI_ISL_592354\|202 | ···················· | ···················· | ···················· | ···················· | ···················· | ···················· |
| M | 6763 | hCoV-19/Australia/VIC14154/2020\|EPI_ISL_592355\|202 | ···················· | ···················· | ···················· | ···················· | ···················· | ···················· |
| M | 6764 | hCoV-19/Australia/VIC14155/2020\|EPI_ISL_592356\|202 | ·····R·············· | ···················· | ···················· | ···················· | ···················· | ···················· |
| M | 6765 | hCoV-19/Australia/VIC14157/2020\|EPI_ISL_592357\|202 | ···················· | ···················· | ···················· | ···················· | ···················· | ···················· |
| M | 6766 | hCoV-19/Australia/VIC14158/2020\|EPI_ISL_592358\|202 | ···················· | ···················· | ···················· | ···················· | ···················· | ···················· |
| M | 6767 | hCoV-19/Australia/VIC14160/2020\|EPI_ISL_592359\|202 | ···················· | ···················· | ···················· | ···················· | ···················· | ···················· |
| M | 6769 | hCoV-19/Australia/VIC14162/2020\|EPI_ISL_592361\|202 | ···················· | ···················· | ···················· | ···················· | ···················· | ···················· |
| M | 6770 | hCoV-19/Australia/VIC14163/2020\|EPI_ISL_592362\|202 | ···················· | ···················· | ···················· | ···················· | ···················· | ···················· |
| M | 6771 | hCoV-19/Australia/VIC14165/2020\|EPI_ISL_592363\|202 | ···················· | ···················· | ···················· | ···················· | ···················· | ···················· |
| M | 6772 | hCoV-19/Australia/VIC14168/2020\|EPI_ISL_592364\|202 | ···················· | ···················· | ···················· | ···················· | ···················· | ···················· |
| M | 6773 | hCoV-19/Australia/VIC14170/2020\|EPI_ISL_592365\|202 | ···················· | ···················· | ···················· | ···················· | ···················· | ···················· |
| M | 6774 | hCoV-19/Australia/VIC14171/2020\|EPI_ISL_592366\|202 | ···················· | ···················· | ···················· | ···················· | ···················· | ···················· |
| M | 6775 | hCoV-19/Australia/VIC14172/2020\|EPI_ISL_592367\|202 | ···················· | ···················· | ···················· | ···················· | ···················· | ···················· |
| M | 6778 | hCoV-19/Australia/VIC14177/2020\|EPI_ISL_592370\|202 | ···················· | ···················· | ···················· | ···················· | ···················· | ···················· |
| M | 6779 | hCoV-19/Australia/VIC14179/2020\|EPI_ISL_592371\|202 | ···················· | ···················· | ···················· | ···················· | ···················· | ···················· |
| M | 6780 | hCoV-19/Australia/VIC14182/2020\|EPI_ISL_592372\|202 | ···················· | ···················· | ···················· | ···················· | ···················· | ···················· |
| M | 6782 | hCoV-19/Australia/VIC14184/2020\|EPI_ISL_592374\|202 | ···················· | ···················· | ···················· | ···················· | ···················· | ···················· |
| M | 6783 | hCoV-19/Australia/VIC14185/2020\|EPI_ISL_592375\|202 | ···················· | ···················· | ···················· | ···················· | ···················· | ···················· |
| M | 6784 | hCoV-19/Australia/VIC14186/2020\|EPI_ISL_592376\|202 | ···················· | ·····A·············· | ···················· | ···················· | ···················· | ···················· |
| M | 6785 | hCoV-19/Australia/VIC14188/2020\|EPI_ISL_592377\|202 | ···················· | ···················· | ···················· | ···················· | ···················· | ···················· |
| M | 6787 | hCoV-19/Australia/VIC14192/2020\|EPI_ISL_592379\|202 | ···················· | ···················· | ···················· | ···················· | ···················· | ···················· |
| M | 6789 | hCoV-19/Australia/VIC14194/2020\|EPI_ISL_592381\|202 | ···················· | ···················· | ···················· | ···················· | ···················· | ···················· |
| M | 6790 | hCoV-19/Australia/VIC14195/2020\|EPI_ISL_592382\|202 | ···················· | ···················· | ···················· | ···················· | ···················· | ···················· |
| M | 6791 | hCoV-19/Australia/VIC14198/2020\|EPI_ISL_592383\|202 | ···················· | ···················· | ···················· | ···················· | ···················· | ···················· |
| M | 6792 | hCoV-19/Australia/VIC14200/2020\|EPI_ISL_592384\|202 | ···················· | ···················· | ···················· | ···················· | ···················· | ···················· |
| M | 6793 | hCoV-19/Australia/QLD1091/2020\|EPI_ISL_593610\|2020 | ···················· | ···················· | ···················· | ···················· | ···················· | ···················· |
| M | 6794 | hCoV-19/Australia/QLD1092/2020\|EPI_ISL_593611\|2020 | ···················· | ···················· | ···················· | ···················· | ···················· | ···················· |
| M | 6795 | hCoV-19/Australia/QLD1093/2020\|EPI_ISL_593612\|2020 | ···················· | ···················· | ···················· | ···················· | ···················· | ···················· |
| M | 6796 | hCoV-19/Australia/QLD1094/2020\|EPI_ISL_593613\|2020 | ···················· | ···················· | ···················· | ···················· | ···················· | ···················· |

| | | | | | | | |
|---|---|---|---|---|---|---|---|
| M | 6797 hCoV-19/Australia/QLD1113/2020\|EPI_ISL_593630\|2020 | ···················· | ···················· | ····················· | ···················· | ···················· | ···················· |
| M | 6798 hCoV-19/Australia/QLD1114/2020\|EPI_ISL_593631\|2020 | ···················· | ···················· | ····················· | ···················· | ···················· | ···················· |
| M | 6799 hCoV-19/Australia/QLD1115/2020\|EPI_ISL_593632\|2020 | ···················· | ···················· | ····················· | ···················· | ···················· | ···················· |
| M | 6800 hCoV-19/Australia/QLD1117/2020\|EPI_ISL_593633\|2020 | ···················· | ···················· | ····················· | ···················· | ···················· | ···················· |
| M | 6801 hCoV-19/Australia/QLD1118/2020\|EPI_ISL_593634\|2020 | ···················· | ···················· | ····················· | ···················· | ···················· | ···················· |
| M | 6802 hCoV-19/Australia/QLD1119/2020\|EPI_ISL_593635\|2020 | ···················· | ···················· | ····················· | ···················· | ···················· | ···················· |
| M | 6803 hCoV-19/Australia/QLD1121/2020\|EPI_ISL_593636\|2020 | ···················· | ···················· | ····················· | ···················· | ···················· | ···················· |
| M | 6804 hCoV-19/Australia/QLD1132/2020\|EPI_ISL_593638\|2020 | ···················· | ···················· | ····················· | ···················· | ···················· | ···················· |
| M | 6805 hCoV-19/Australia/QLD1134/2020\|EPI_ISL_593639\|2020 | ···················· | ···················· | ····················· | ···················· | ···················· | ···················· |
| M | 6806 hCoV-19/Australia/QLD1138/2020\|EPI_ISL_593640\|2020 | ···················· | ···················· | ····················· | ···················· | ···················· | ···················· |
| M | 6807 hCoV-19/Australia/QLD1140/2020\|EPI_ISL_593641\|2020 | ···················· | ···················· | ····················· | ···················· | ···················· | ···················· |
| M | 6808 hCoV-19/Australia/QLD1141/2020\|EPI_ISL_593642\|2020 | ···················· | ···················· | ····················· | ···················· | ···················· | ···················· |
| M | 6809 hCoV-19/Australia/QLD1142/2020\|EPI_ISL_593643\|2020 | ···················· | ···················· | ····················· | ···················· | ···················· | ···················· |
| M | 6810 hCoV-19/Australia/QLD1143/2020\|EPI_ISL_593644\|2020 | ···················· | ···················· | ····················· | ···················· | ···················· | ···················· |
| M | 6811 hCoV-19/Australia/QLD1146/2020\|EPI_ISL_593645\|2020 | ···················· | ···················· | ····················· | ···················· | ···················· | ···················· |
| M | 6812 hCoV-19/Australia/NSW1058/2020\|EPI_ISL_593646\|2020 | ···················· | ···················· | ····················· | ···················· | ···················· | ···················· |
| M | 6813 hCoV-19/Australia/NSW1091/2020\|EPI_ISL_593647\|2020 | ···················· | ···················· | ····················· | ···················· | ···················· | ···················· |
| M | 6814 hCoV-19/Australia/NSW1092/2020\|EPI_ISL_593648\|2020 | ···················· | ···················· | ····················· | ···················· | ···················· | ···················· |
| M | 6815 hCoV-19/Australia/NSW1060/2020\|EPI_ISL_593649\|2020 | ···················· | ···················· | ····················· | ···················· | ···················· | ···················· |
| M | 6816 hCoV-19/Australia/NSW1061/2020\|EPI_ISL_593650\|2020 | ···················· | ···················· | ····················· | ···················· | ···················· | ···················· |
| M | 6817 hCoV-19/Australia/NSW1088/2020\|EPI_ISL_593651\|2020 | ···················· | ···················· | ····················· | ···················· | ···················· | ···················· |
| M | 6818 hCoV-19/Australia/NSW1095/2020\|EPI_ISL_593652\|2020 | ···················· | ···················· | ····················· | ···················· | ···················· | ···················· |
| M | 6819 hCoV-19/Australia/NSW1080/2020\|EPI_ISL_593653\|2020 | ···················· | ···················· | ····················· | ···················· | ···················· | ···················· |
| M | 6820 hCoV-19/Australia/NSW1056/2020\|EPI_ISL_593655\|2020 | ···················· | ···················· | ····················· | ···················· | ···················· | ···················· |
| M | 6821 hCoV-19/Australia/NSW1096/2020\|EPI_ISL_593658\|2020 | ···················· | ···················· | ····················· | ···················· | ···················· | ···················· |
| M | 6822 hCoV-19/Australia/NSW1071/2020\|EPI_ISL_593659\|2020 | ···················· | ···················· | ····················· | ···················· | ···················· | ···················· |
| M | 6823 hCoV-19/Australia/NSW1016/2020\|EPI_ISL_593661\|2020 | ···················· | ···················· | ····················· | ···················· | ···················· | ···················· |
| M | 6824 hCoV-19/Australia/NSW1017/2020\|EPI_ISL_593662\|2020 | ···················· | ···················· | ····················· | ···················· | ···················· | ···················· |
| M | 6825 hCoV-19/Australia/NSW1019/2020\|EPI_ISL_593664\|2020 | ···················· | ···················· | ····················· | ···················· | ···················· | ···················· |
| M | 6826 hCoV-19/Australia/NSW1020/2020\|EPI_ISL_593665\|2020 | ···················· | ···················· | ····················· | ···················· | ···················· | ···················· |
| M | 6827 hCoV-19/Australia/NSW1023/2020\|EPI_ISL_593667\|2020 | ···················· | ···················· | ····················· | ···················· | ···················· | ···················· |
| M | 6828 hCoV-19/Australia/NSW1031/2020\|EPI_ISL_593668\|2020 | ···················· | ···················· | ····················· | ···················· | ···················· | ···················· |
| M | 6829 hCoV-19/Australia/NSW1057/2020\|EPI_ISL_593669\|2020 | ···················· | ···················· | ····················· | ···················· | ···················· | ···················· |
| M | 6830 hCoV-19/Australia/NSW1059/2020\|EPI_ISL_593670\|2020 | ···················· | ···················· | ····················· | ···················· | ···················· | ···················· |
| M | 6831 hCoV-19/Australia/NSW1062/2020\|EPI_ISL_593671\|2020 | ···················· | ···················· | ····················· | ···················· | ···················· | ···················· |
| M | 6832 hCoV-19/Australia/NSW1063/2020\|EPI_ISL_593672\|2020 | ···················· | ···················· | ····················· | ···················· | ···················· | ···················· |
| M | 6833 hCoV-19/Australia/NSW1065/2020\|EPI_ISL_593673\|2020 | ···················· | ···················· | ····················· | ···················· | ···················· | ···················· |
| M | 6834 hCoV-19/Australia/NSW1066/2020\|EPI_ISL_593674\|2020 | ···················· | ···················· | ····················· | ···················· | ···················· | ···················· |
| M | 6835 hCoV-19/Australia/NSW1067/2020\|EPI_ISL_593675\|2020 | ···················· | ···················· | ····················· | ···················· | ···················· | ···················· |
| M | 6836 hCoV-19/Australia/NSW1074/2020\|EPI_ISL_593676\|2020 | ···················· | ···················· | ····················· | ···················· | ···················· | ···················· |
| M | 6837 hCoV-19/Australia/NSW1075/2020\|EPI_ISL_593677\|2020 | ···················· | ···················· | ····················· | ···················· | ···················· | ···················· |
| M | 6838 hCoV-19/Australia/NSW1082/2020\|EPI_ISL_593678\|2020 | ···················· | ···················· | ····················· | ···················· | ···················· | ···················· |
| M | 6839 hCoV-19/Australia/NSW1084/2020\|EPI_ISL_593679\|2020 | ···················· | ···················· | ····················· | ···················· | ···················· | ···················· |
| M | 6840 hCoV-19/Australia/NSW1085/2020\|EPI_ISL_593680\|2020 | ···················· | ···················· | ····················· | ···················· | ···················· | ···················· |
| M | 6841 hCoV-19/Australia/NSW1087/2020\|EPI_ISL_593681\|2020 | ···················· | ···················· | ····················· | ···················· | ···················· | ···················· |
| M | 6842 hCoV-19/Australia/NSW1093/2020\|EPI_ISL_593682\|2020 | ···················· | ···················· | ····················· | ···················· | ···················· | ···················· |
| M | 6843 hCoV-19/Australia/NSW1026/2020\|EPI_ISL_593683\|2020 | ···················· | ···················· | ····················· | ···················· | ···················· | ···················· |
| M | 6844 hCoV-19/Australia/NSW1032/2020\|EPI_ISL_593684\|2020 | ···················· | ···················· | ····················· | ···················· | ···················· | ···················· |
| M | 6845 hCoV-19/Australia/NSW1033/2020\|EPI_ISL_593685\|2020 | ···················· | ···················· | ····················· | ···················· | ···················· | ···················· |
| M | 6846 hCoV-19/Australia/NSW1034/2020\|EPI_ISL_593686\|2020 | ···················· | ···················· | ····················· | ···················· | ···················· | ···················· |
| M | 6847 hCoV-19/Australia/NSW1035/2020\|EPI_ISL_593687\|2020 | ···················· | ···················· | ····················· | ···················· | ···················· | ···················· |
| M | 6848 hCoV-19/Australia/NSW1036/2020\|EPI_ISL_593688\|2020 | ···················· | ·A·················· | ····················· | ···················· | ···················· | ··A················· |
| M | 6849 hCoV-19/Australia/NSW1037/2020\|EPI_ISL_593689\|2020 | ···················· | ···················· | ····················· | ···················· | ···················· | ···················· |
| M | 6850 hCoV-19/Australia/NSW1038/2020\|EPI_ISL_593690\|2020 | ···················· | ···················· | ····················· | ···················· | ···················· | ···················· |
| M | 6851 hCoV-19/Australia/NSW1039/2020\|EPI_ISL_593691\|2020 | ···················· | ···················· | ····················· | ···················· | ···················· | ···················· |
| M | 6852 hCoV-19/Australia/NSW1040/2020\|EPI_ISL_593692\|2020 | ···················· | ···················· | ····················· | ···················· | ···················· | ···················· |
| M | 6853 hCoV-19/Australia/NSW1041/2020\|EPI_ISL_593693\|2020 | ···················· | ···················· | ····················· | ···················· | ···················· | ···················· |
| M | 6854 hCoV-19/Australia/NSW1042/2020\|EPI_ISL_593694\|2020 | ···················· | ···················· | ····················· | ···················· | ···················· | ···················· |
| M | 6855 hCoV-19/Australia/NSW1043/2020\|EPI_ISL_593695\|2020 | ···················· | ···················· | ····················· | ···················· | ···················· | ···················· |
| M | 6856 hCoV-19/Australia/NSW1044/2020\|EPI_ISL_593696\|2020 | ···················· | ···················· | ····················· | ···················· | ···················· | ···················· |
| M | 6857 hCoV-19/Australia/NSW1045/2020\|EPI_ISL_593697\|2020 | ···················· | ···················· | ····················· | ···················· | ···················· | ···················· |
| M | 6858 hCoV-19/Australia/NSW1046/2020\|EPI_ISL_593698\|2020 | ···················· | ···················· | ····················· | ···················· | ···················· | ···················· |
| M | 6859 hCoV-19/Australia/NSW1047/2020\|EPI_ISL_593699\|2020 | ···················· | ···················· | ····················· | ···················· | ···················· | ···················· |
| M | 6860 hCoV-19/England/MILK-9EF0E0/2020\|EPI_ISL_594900\|20 | ···················· | ·A·················· | ····················· | ···················· | ···················· | ··A················· |
| M | 6861 hCoV-19/England/MILK-9EF10B/2020\|EPI_ISL_594901\|20 | ···················· | ·A·················· | ····················· | ···················· | ···················· | ··A················· |
| M | 6862 hCoV-19/Wales/MILK-9EF147/2020\|EPI_ISL_594902\|2020 | ···················· | ···················· | ····················· | ···················· | ···················· | ···················· |
| M | 6863 hCoV-19/England/MILK-9EF156/2020\|EPI_ISL_594903\|20 | ···················· | ···················· | ····················· | ···················· | ···················· | ···················· |
| M | 6864 hCoV-19/Wales/MILK-9EF165/2020\|EPI_ISL_594904\|2020 | ···················· | ···················· | ····················· | ···················· | ···················· | ···················· |
| M | 6865 hCoV-19/Wales/MILK-9EF183/2020\|EPI_ISL_594905\|2020 | ···················· | ···················· | ····················· | ···················· | ···················· | ···················· |
| M | 6866 hCoV-19/Wales/MILK-9EF192/2020\|EPI_ISL_594906\|2020 | ···················· | ···················· | ····················· | ···················· | ···················· | ···················· |
| M | 6867 hCoV-19/England/MILK-9EF1DE/2020\|EPI_ISL_594907\|20 | ···················· | ···················· | ····················· | ···················· | ···················· | ···················· |
| M | 6868 hCoV-19/England/MILK-9EF1FC/2020\|EPI_ISL_594908\|20 | ···················· | ·A·················· | ····················· | ···················· | ···················· | ··A················· |
| M | 6869 hCoV-19/England/MILK-9EF235/2020\|EPI_ISL_594909\|20 | ···················· | ·A·················· | ····················· | ···················· | ···················· | ··A················· |
| M | 6870 hCoV-19/England/MILK-9EF244/2020\|EPI_ISL_594910\|20 | ···················· | ···················· | ····················· | ···················· | ···················· | ···················· |
| M | 6871 hCoV-19/England/MILK-9EF262/2020\|EPI_ISL_594911\|20 | ···················· | ···················· | ····················· | ···················· | ···················· | ···················· |
| M | 6872 hCoV-19/England/MILK-9EF280/2020\|EPI_ISL_594912\|20 | ···················· | ···················· | ····················· | ···················· | ···················· | ···················· |
| M | 6873 hCoV-19/England/MILK-9EF314/2020\|EPI_ISL_594913\|20 | ···················· | ···················· | ····················· | ···················· | ···················· | ···················· |
| M | 6874 hCoV-19/England/MILK-9EF38D/2020\|EPI_ISL_594914\|20 | ···················· | ···················· | ····················· | ···················· | ···················· | ···················· |
| M | 6875 hCoV-19/Wales/MILK-9EF420/2020\|EPI_ISL_594916\|2020 | ···················· | ···················· | ····················· | ···················· | ···················· | ···················· |
| M | 6876 hCoV-19/England/MILK-9EF44E/2020\|EPI_ISL_594917\|20 | ···················· | ···················· | ····················· | ···················· | ···················· | ···················· |

| | | | | | | | |
|---|---|---|---|---|---|---|---|
| M | 6877 hCoV-19/England/MILK-9EF47B/2020\|EPI_ISL_594918\|20 | ···················· | ···················· | ···················· | ···················· | ···················· | ···················· |
| M | 6878 hCoV-19/Wales/MILK-9EF499/2020\|EPI_ISL_594919\|2020 | ···················· | ···················· | ···················· | ···················· | ···················· | ···················· |
| M | 6879 hCoV-19/England/MILK-9EF7BE/2020\|EPI_ISL_594941\|20 | ···················· | ···················· | ···················· | ···················· | ···················· | ···················· |
| M | 6880 hCoV-19/England/MILK-9EF7CD/2020\|EPI_ISL_594942\|20 | ···················· | ···················· | ···················· | ···················· | ···················· | ···················· |
| M | 6881 hCoV-19/England/MILK-9EF7DC/2020\|EPI_ISL_594943\|20 | ···················· | ···················· | ···················· | ···················· | ···················· | ···················· |
| M | 6882 hCoV-19/England/MILK-9EF7FA/2020\|EPI_ISL_594944\|20 | ···················· | ···················· | ···················· | ···················· | ···················· | ···················· |
| M | 6883 hCoV-19/England/MILK-9EF96D/2020\|EPI_ISL_594961\|20 | ···················· | ···················· | ···················· | ···················· | ···················· | ···················· |
| M | 6884 hCoV-19/England/MILK-9EF97C/2020\|EPI_ISL_594962\|20 | ···················· | ···················· | ···················· | ···················· | ···················· | ···················· |
| M | 6885 hCoV-19/Wales/MILK-9EF9A9/2020\|EPI_ISL_594963\|2020 | ···················· | ···················· | ···················· | ···················· | ···················· | ···················· |
| M | 6886 hCoV-19/England/MILK-9EF9B8/2020\|EPI_ISL_594964\|20 | ···················· | ···················· | ···················· | ···················· | ···················· | ···················· |
| M | 6887 hCoV-19/England/MILK-9EF9D6/2020\|EPI_ISL_594965\|20 | ···················· | ···················· | ···················· | ···················· | ···················· | ···················· |
| M | 6888 hCoV-19/Wales/MILK-9EF9E5/2020\|EPI_ISL_594966\|2020 | ···················· | ···················· | ···················· | ···················· | ···················· | ···················· |
| M | 6889 hCoV-19/Wales/MILK-9EF9F4/2020\|EPI_ISL_594967\|2020 | ···················· | ···················· | ···················· | ···················· | ···················· | ···················· |
| M | 6890 hCoV-19/Wales/MILK-9EFA00/2020\|EPI_ISL_594968\|2020 | ···················· | ···················· | ···················· | ···················· | ···················· | ···················· |
| M | 6891 hCoV-19/Wales/MILK-9EFA1F/2020\|EPI_ISL_594969\|2020 | ···················· | ···················· | ···················· | ···················· | ···················· | ···················· |
| M | 6892 hCoV-19/Wales/MILK-9EFA2E/2020\|EPI_ISL_594970\|2020 | ···················· | ···················· | ···················· | ···················· | ···················· | ···················· |
| M | 6893 hCoV-19/Wales/MILK-9EFA3D/2020\|EPI_ISL_594971\|2020 | ···················· | ···················· | ···················· | ···················· | ···················· | ···················· |
| M | 6894 hCoV-19/Wales/MILK-9EFA4C/2020\|EPI_ISL_594972\|2020 | ···················· | ···················· | ···················· | ···················· | ···················· | ···················· |
| M | 6895 hCoV-19/Wales/MILK-9EFA6A/2020\|EPI_ISL_594973\|2020 | ···················· | ···················· | ···················· | ···················· | ···················· | ···················· |
| M | 6896 hCoV-19/England/MILK-9EFA88/2020\|EPI_ISL_594974\|20 | ···················· | ···················· | ···················· | ···················· | ···················· | ···················· |
| M | 6897 hCoV-19/Wales/MILK-9EFA97/2020\|EPI_ISL_594975\|2020 | ···················· | ···················· | ···················· | ···················· | ···················· | ···················· |
| M | 6898 hCoV-19/England/MILK-9EFAB5/2020\|EPI_ISL_594976\|20 | ···················· | ···················· | ···················· | ···················· | ···················· | ···················· |
| M | 6899 hCoV-19/Wales/MILK-9EFAC4/2020\|EPI_ISL_594977\|2020 | ···················· | ···················· | ···················· | ···················· | ···················· | ···················· |
| M | 6900 hCoV-19/Wales/MILK-9EFAD3/2020\|EPI_ISL_594978\|2020 | ···················· | ···················· | ···················· | ···················· | ···················· | ···················· |
| M | 6901 hCoV-19/England/MILK-9EFAE2/2020\|EPI_ISL_594979\|20 | ···················· | ···················· | ···················· | ···················· | ···················· | ···················· |
| M | 6902 hCoV-19/England/MILK-9EFAF1/2020\|EPI_ISL_594980\|20 | ···················· | ·A·················· | ···················· | ···················· | ···················· | ··A················· |
| M | 6903 hCoV-19/Wales/MILK-9EFB49/2020\|EPI_ISL_594983\|2020 | ···················· | ···················· | ···················· | ···················· | ···················· | ···················· |
| M | 6904 hCoV-19/England/MILK-9EFBB2/2020\|EPI_ISL_594985\|20 | ···················· | ·A·················· | ···················· | ···················· | ···················· | ··A················· |
| M | 6905 hCoV-19/England/MILK-A17DE7/2020\|EPI_ISL_594989\|20 | ···················· | ···················· | ···················· | ···················· | ···················· | ···················· |
| M | 6906 hCoV-19/England/MILK-A17E3F/2020\|EPI_ISL_594992\|20 | ···················· | ···················· | ···················· | ···················· | ···················· | ···················· |
| M | 6907 hCoV-19/England/MILK-A17E4E/2020\|EPI_ISL_594993\|20 | ···················· | ···················· | ···················· | ···················· | ···················· | ···················· |
| M | 6908 hCoV-19/England/MILK-A17E7B/2020\|EPI_ISL_594994\|20 | ···················· | ···················· | ···················· | ···················· | ···················· | ···················· |
| M | 6909 hCoV-19/England/MILK-A17E99/2020\|EPI_ISL_594995\|20 | ···················· | ···················· | ···················· | ···················· | ···················· | ···················· |
| M | 6910 hCoV-19/England/MILK-A17EE4/2020\|EPI_ISL_594997\|20 | ···················· | ···················· | ···················· | ···················· | ···················· | ···················· |
| M | 6911 hCoV-19/England/MILK-A17EF3/2020\|EPI_ISL_594998\|20 | ···················· | ···················· | ···················· | ···················· | ···················· | ···················· |
| M | 6912 hCoV-19/England/MILK-A17F0F/2020\|EPI_ISL_594999\|20 | ···················· | ···················· | ···················· | ···················· | ···················· | ···················· |
| M | 6913 hCoV-19/Northern Ireland/NIRE-10876C/2020\|EPI_ISL_ | ···················· | ···················· | ···················· | ···················· | ···················· | ···················· |
| M | 6914 hCoV-19/Northern Ireland/NIRE-10877B/2020\|EPI_ISL_ | ···················· | ···················· | ···················· | ···················· | ···················· | ···················· |
| M | 6915 hCoV-19/Northern Ireland/NIRE-1087A8/2020\|EPI_ISL_ | ···················· | ···················· | ···················· | ···················· | ···················· | ···················· |
| M | 6916 hCoV-19/Northern Ireland/NIRE-1088B4/2020\|EPI_ISL_ | ···················· | ···················· | ···················· | ···················· | ···················· | ···················· |
| M | 6917 hCoV-19/Northern Ireland/NIRE-1088C3/2020\|EPI_ISL_ | ···················· | ···················· | ···················· | ···················· | ···················· | ···················· |
| M | 6918 hCoV-19/Northern Ireland/NIRE-1088D2/2020\|EPI_ISL_ | ···················· | ···················· | ···················· | ···················· | ···················· | ···················· |
| M | 6919 hCoV-19/Northern Ireland/NIRE-1088E1/2020\|EPI_ISL_ | ···················· | ···················· | ···················· | ···················· | ···················· | ···················· |
| M | 6920 hCoV-19/Northern Ireland/NIRE-10892A/2020\|EPI_ISL_ | ···················· | ···················· | ···················· | ···················· | ···················· | ···················· |
| M | 6921 hCoV-19/Northern Ireland/NIRE-108939/2020\|EPI_ISL_ | ···················· | ···················· | ···················· | ···················· | ···················· | ···················· |
| M | 6922 hCoV-19/Northern Ireland/NIRE-108948/2020\|EPI_ISL_ | ···················· | ···················· | ···················· | ···················· | ···················· | ···················· |
| M | 6923 hCoV-19/Northern Ireland/NIRE-108984/2020\|EPI_ISL_ | ···················· | ···················· | ···················· | ···················· | ···················· | NN·················· |
| M | 6924 hCoV-19/Northern Ireland/NIRE-FB2A8/2020\|EPI_ISL_5 | ···················· | ···················· | ···················· | ···················· | ···················· | ···················· |
| M | 6925 hCoV-19/Northern Ireland/NIRE-FB2C6/2020\|EPI_ISL_5 | ···················· | ···················· | ···················· | ···················· | ···················· | ···················· |
| M | 6926 hCoV-19/Northern Ireland/NIRE-FB2E4/2020\|EPI_ISL_5 | ···················· | ···················· | ···················· | ···················· | ···················· | ···················· |
| M | 6927 hCoV-19/Northern Ireland/NIRE-FB30F/2020\|EPI_ISL_5 | ···················· | ···················· | ···················· | ···················· | ···················· | ···················· |
| M | 6928 hCoV-19/Northern Ireland/NIRE-FB34B/2020\|EPI_ISL_5 | ···················· | ···················· | ···················· | ···················· | ···················· | ···················· |
| M | 6930 hCoV-19/Northern Ireland/NIRE-FB387/2020\|EPI_ISL_5 | ···················· | ···················· | ···················· | ···················· | ···················· | ···················· |
| M | 6932 hCoV-19/Northern Ireland/NIRE-FB518/2020\|EPI_ISL_5 | ···················· | ···················· | ···················· | ···················· | ···················· | ···················· |
| M | 6933 hCoV-19/Northern Ireland/NIRE-FB527/2020\|EPI_ISL_5 | ···················· | ···················· | ···················· | ···················· | ···················· | ···················· |
| M | 6935 hCoV-19/Northern Ireland/NIRE-FB545/2020\|EPI_ISL_5 | ···················· | ···················· | ···················· | ···················· | ···················· | ···················· |
| M | 6937 hCoV-19/Northern Ireland/NIRE-FB563/2020\|EPI_ISL_5 | ···················· | ···················· | ···················· | ···················· | ···················· | ···················· |
| M | 6938 hCoV-19/Northern Ireland/NIRE-FB572/2020\|EPI_ISL_5 | ···················· | ···················· | ···················· | ···················· | ···················· | ···················· |
| M | 6939 hCoV-19/Northern Ireland/NIRE-FB581/2020\|EPI_ISL_5 | ···················· | ···················· | ···················· | ···················· | ···················· | ···················· |
| M | 6941 hCoV-19/Northern Ireland/NIRE-FB5BE/2020\|EPI_ISL_5 | ···················· | ···················· | ···················· | ···················· | ···················· | ···················· |
| M | 6942 hCoV-19/Northern Ireland/NIRE-FB5DC/2020\|EPI_ISL_5 | ···················· | ···················· | ···················· | ···················· | ···················· | ···················· |
| M | 6943 hCoV-19/Northern Ireland/NIRE-FB5FA/2020\|EPI_ISL_5 | ···················· | ···················· | ···················· | ···················· | ···················· | ···················· |
| M | 6944 hCoV-19/Northern Ireland/NIRE-FB606/2020\|EPI_ISL_5 | ···················· | ···················· | ···················· | ···················· | ···················· | ···················· |
| M | 6945 hCoV-19/Northern Ireland/NIRE-FB624/2020\|EPI_ISL_5 | ···················· | ···················· | ···················· | ···················· | ···················· | ···················· |
| M | 6946 hCoV-19/Northern Ireland/NIRE-FB642/2020\|EPI_ISL_5 | ···················· | ···················· | ···················· | ···················· | ···················· | ···················· |
| M | 6948 hCoV-19/Northern Ireland/NIRE-FB8E2/2020\|EPI_ISL_5 | ···················· | ···················· | ···················· | ···················· | ···················· | ···················· |
| M | 6949 hCoV-19/Northern Ireland/NIRE-FB91C/2020\|EPI_ISL_5 | ···················· | ···················· | ···················· | ···················· | ···················· | ···················· |
| M | 6950 hCoV-19/Northern Ireland/NIRE-FB92B/2020\|EPI_ISL_5 | ···················· | ···················· | ···················· | ···················· | ···················· | ···················· |
| M | 6951 hCoV-19/Northern Ireland/NIRE-FB93A/2020\|EPI_ISL_5 | ···················· | ···················· | ···················· | ···················· | ···················· | ···················· |
| M | 6952 hCoV-19/Northern Ireland/NIRE-FB949/2020\|EPI_ISL_5 | ···················· | ···················· | ···················· | ···················· | ···················· | ···················· |
| M | 6953 hCoV-19/Northern Ireland/NIRE-FB958/2020\|EPI_ISL_5 | ···················· | ···················· | ···················· | ···················· | ···················· | ···················· |
| M | 6954 hCoV-19/Northern Ireland/NIRE-FB967/2020\|EPI_ISL_5 | ···················· | ···················· | ···················· | ···················· | ···················· | ···················· |
| M | 6956 hCoV-19/Northern Ireland/NIRE-FB985/2020\|EPI_ISL_5 | ···················· | ···················· | ···················· | ···················· | ···················· | ···················· |
| M | 6957 hCoV-19/Northern Ireland/NIRE-FB9B2/2020\|EPI_ISL_5 | ···················· | ···················· | ···················· | ···················· | ···················· | ···················· |
| M | 6958 hCoV-19/Northern Ireland/NIRE-FB9C1/2020\|EPI_ISL_5 | ···················· | ···················· | ···················· | ···················· | ···················· | ···················· |
| M | 6959 hCoV-19/Scotland/CVR4384/2020\|EPI_ISL_585261\|2020- | ···················· | ···················· | ···················· | ···················· | ···················· | ···················· |
| M | 6960 hCoV-19/Scotland/CVR4389/2020\|EPI_ISL_585263\|2020- | ···················· | ···················· | ···················· | ···················· | ···················· | ···················· |
| M | 6961 hCoV-19/Scotland/CVR4393/2020\|EPI_ISL_585267\|2020- | ···················· | ···················· | ···················· | ···················· | ···················· | ···················· |
| M | 6962 hCoV-19/Scotland/CVR4394/2020\|EPI_ISL_585268\|2020- | ···················· | ···················· | ···················· | ···················· | ···················· | ···················· |
| M | 6963 hCoV-19/Scotland/CVR4396/2020\|EPI_ISL_585270\|2020- | ···················· | ···················· | ···················· | ···················· | ···················· | ···················· |

M 6964 hCoV-19/Scotland/CVR4398/2020|EPI_ISL_585272|2020- ···················· ···················· ···················· ···················· ···················· ····················
M 6965 hCoV-19/Scotland/CVR4402/2020|EPI_ISL_585276|2020- ···················· ···················· ···················· ···················· ···················· ····················
M 6966 hCoV-19/Scotland/CVR4403/2020|EPI_ISL_585277|2020- ···················· ···················· ···················· ···················· ···················· ····················
M 6967 hCoV-19/Scotland/CVR4407/2020|EPI_ISL_585281|2020- ···················· ···················· ···················· ···················· ···················· ····················
M 6968 hCoV-19/Scotland/CVR4408/2020|EPI_ISL_585282|2020- ···················· ···················· ···················· ···················· ···················· ····················
M 6969 hCoV-19/Scotland/CVR4410/2020|EPI_ISL_585284|2020- ···················· ···················· ···················· ···················· ···················· ····················
M 6970 hCoV-19/Scotland/CVR4411/2020|EPI_ISL_585285|2020- ···················· ·A·················· ···················· ···················· ···················· ····················
M 6971 hCoV-19/Scotland/CVR4412/2020|EPI_ISL_585286|2020- ···················· ···················· ···················· ···················· ···················· ····················
M 6972 hCoV-19/Scotland/CVR4413/2020|EPI_ISL_585287|2020- ···················· ···················· ···················· ···················· ···················· ····················
M 6973 hCoV-19/Scotland/CVR4414/2020|EPI_ISL_585288|2020- ···················· ···················· ···················· ···················· ···················· ····················
M 6974 hCoV-19/Scotland/CVR4416/2020|EPI_ISL_585290|2020- ···················· ···················· ···················· ···················· ···················· ····················
M 6975 hCoV-19/Scotland/CVR4417/2020|EPI_ISL_585291|2020- ···················· ···················· ···················· ···················· ···················· ····················
M 6976 hCoV-19/Scotland/CVR4419/2020|EPI_ISL_585293|2020- ···················· ···················· ···················· ···················· ···················· ····················
M 6977 hCoV-19/Scotland/CVR4421/2020|EPI_ISL_585295|2020- ···················· ···················· ···················· ···················· ···················· ····················
M 6978 hCoV-19/Scotland/CVR4425/2020|EPI_ISL_585297|2020- ···················· ···················· ···················· ···················· ···················· ····················
M 6979 hCoV-19/Scotland/CVR4428/2020|EPI_ISL_585299|2020- ···················· ···················· ···················· ···················· ···················· ····················
M 6980 hCoV-19/India/GJ-GBRC-403a/2020|EPI_ISL_586522|202 ···················· ·A·················· ···················· ···················· ···················· ··A·················
M 6981 hCoV-19/India/GJ-GBRC-403b/2020|EPI_ISL_586523|202 ···················· ·A·················· ···················· ···················· ···················· ··A·················
M 6982 hCoV-19/India/GJ-GBRC-404a/2020|EPI_ISL_586524|202 ···················· ·A·················· ···················· ···················· ···················· ··A·················
M 6983 hCoV-19/India/GJ-GBRC-404b/2020|EPI_ISL_586525|202 ···················· ·A·················· ···················· ···················· ···················· ··A·················
M 6984 hCoV-19/India/GJ-GBRC-405a/2020|EPI_ISL_586526|202 ···················· ···················· ···················· ···················· ···················· ····················
M 6985 hCoV-19/India/GJ-GBRC-405b/2020|EPI_ISL_586527|202 ···················· ·A·················· ···················· ···················· ···················· ··A·················
M 6986 hCoV-19/India/GJ-GBRC-406b/2020|EPI_ISL_586528|202 ···················· ·A·················· ···················· ···················· ···················· ··A·················
M 6987 hCoV-19/India/GJ-GBRC-407a/2020|EPI_ISL_586529|202 ···················· ·A·················· ···················· ···················· ···················· ··A·················
M 6988 hCoV-19/India/GJ-GBRC-407b/2020|EPI_ISL_586530|202 ···················· ·A·················· ···················· ···················· ···················· ··A·················
M 6989 hCoV-19/India/GJ-GBRC-408a/2020|EPI_ISL_586531|202 ···················· ···················· ···················· ···················· ···················· ····················
M 6990 hCoV-19/India/GJ-GBRC-408b/2020|EPI_ISL_586532|202 ···················· ·A·················· ···················· ···················· ···················· ····················
M 6991 hCoV-19/India/GJ-GBRC-409b/2020|EPI_ISL_586534|202 ···················· ···················· ···················· ···················· ···················· ····················
M 6992 hCoV-19/India/GJ-GBRC-416a/2020|EPI_ISL_586544|202 ···················· ·A·················· ···················· ···················· ···················· ··A·················
M 6993 hCoV-19/India/GJ-GBRC-416b/2020|EPI_ISL_586545|202 ···················· ·A·················· ···················· ···················· ···················· ··A·················
M 6994 hCoV-19/India/GJ-GBRC-417a/2020|EPI_ISL_586546|202 ···················· ·A·················· ···················· ···················· ···················· ··A·················
M 6995 hCoV-19/India/GJ-GBRC-417b/2020|EPI_ISL_586547|202 ···················· ·A·················· ···················· ···················· ···················· ····················
M 6996 hCoV-19/India/GJ-GBRC-418/2020|EPI_ISL_586548|2020 ···················· ···················· ···················· ···················· ···················· ····················
M 6997 hCoV-19/India/GJ-GBRC-419a/2020|EPI_ISL_586549|202 ···················· ···················· ···················· ···················· ···················· ····················
M 6998 hCoV-19/India/GJ-GBRC-419b/2020|EPI_ISL_586550|202 ···················· ···················· ···················· ···················· ···················· ····················
M 6999 hCoV-19/India/GJ-GBRC-422a/2020|EPI_ISL_586551|202 ···················· ···················· ···················· ···················· ···················· ····················
M 7000 hCoV-19/India/GJ-GBRC-422b/2020|EPI_ISL_586552|202 ···················· ···················· ···················· ···················· ···················· ····················
M 7001 hCoV-19/India/GJ-GBRC-423a/2020|EPI_ISL_586553|202 ···················· ·A·················· ···················· ···················· ···················· ····················
M 7002 hCoV-19/India/GJ-GBRC-423b/2020|EPI_ISL_586554|202 ···················· ···················· ···················· ···················· ···················· ··A·················
M 7003 hCoV-19/India/GJ-GBRC-424a/2020|EPI_ISL_586555|202 ···················· ·A·················· ···················· ···················· ···················· ··A·················
M 7004 hCoV-19/India/GJ-GBRC-424b/2020|EPI_ISL_586556|202 ···················· ·A·················· ···················· ···················· ···················· ··A·················
M 7005 hCoV-19/India/GJ-GBRC-425/2020|EPI_ISL_586557|2020 ···················· ···················· ···················· ···················· ···················· ····················
M 7006 hCoV-19/India/GJ-GBRC-426a/2020|EPI_ISL_586558|202 ···················· ···················· ···················· ···················· ···················· ····················
M 7007 hCoV-19/India/GJ-GBRC-426b/2020|EPI_ISL_586559|202 ···················· ···················· ···················· ···················· ···················· ····················
M 7008 hCoV-19/India/GJ-GBRC-427a/2020|EPI_ISL_586560|202 ···················· ·A·················· ···················· ···················· ···················· ··A·················
M 7009 hCoV-19/India/GJ-GBRC-427b/2020|EPI_ISL_586561|202 ···················· ·A·················· ···················· ···················· ···················· ··A·················
M 7010 hCoV-19/India/GJ-GBRC-428a/2020|EPI_ISL_586562|202 ···················· ·A·················· ···················· ···················· ···················· ··A·················
M 7011 hCoV-19/India/GJ-GBRC-428b/2020|EPI_ISL_586563|202 ···················· ·A·················· ···················· ···················· ···················· ··A·················
M 7012 hCoV-19/India/GJ-GBRC-429/2020|EPI_ISL_586564|2020 ···················· ···················· ···················· ···················· ···················· ····················
M 7013 hCoV-19/India/GJ-GBRC-430a/2020|EPI_ISL_586565|202 ···················· ·A·················· ···················· ···················· ···················· ··A·················
M 7014 hCoV-19/India/GJ-GBRC-430b/2020|EPI_ISL_586566|202 ···················· ·A·················· ···················· ···················· ···················· ··A·················
M 7015 hCoV-19/India/GJ-GBRC-431a/2020|EPI_ISL_586567|202 ···················· ·A·················· ···················· ···················· ···················· ··A·················
M 7016 hCoV-19/India/GJ-GBRC-431b/2020|EPI_ISL_586568|202 ···················· ·A·················· ···················· ···················· ···················· ··A·················
M 7017 hCoV-19/India/GJ-GBRC-432/2020|EPI_ISL_586569|2020 ···················· ···················· ···················· ···················· ···················· ····················
M 7018 hCoV-19/India/GJ-GBRC-433/2020|EPI_ISL_586570|2020 ···················· ···················· ···················· ···················· ···················· ····················
M 7019 hCoV-19/Australia/NSW2881/2020|EPI_ISL_586571|2020 ···················· ···················· ···················· ···················· ···················· ····················
M 7020 hCoV-19/England/ALDP-A2DB99/2020|EPI_ISL_586574|20 ···················· ···················· ···················· ···················· ···················· ····················
M 7021 hCoV-19/England/ALDP-A2DCA5/2020|EPI_ISL_586575|20 ···················· ···················· ···················· ···················· ···················· ····················
M 7022 hCoV-19/England/ALDP-A2DA41/2020|EPI_ISL_586576|20 ···················· ·A·················· ···················· ···················· ···················· ··A·················
M 7023 hCoV-19/England/ALDP-A2E55E/2020|EPI_ISL_586577|20 ···················· ···················· ···················· ···················· ···················· ····················
M 7024 hCoV-19/England/ALDP-A2E56D/2020|EPI_ISL_586578|20 ···················· ···················· ···················· ···················· ···················· ····················
M 7025 hCoV-19/England/ALDP-A2E62E/2020|EPI_ISL_586579|20 ···················· ···················· ···················· ···················· ···················· ····················
M 7026 hCoV-19/England/ALDP-A2D21C/2020|EPI_ISL_586580|20 ···················· ···················· ···················· ···················· ···················· ····················
M 7027 hCoV-19/England/ALDP-A2D67A/2020|EPI_ISL_586581|20 ···················· ···················· ···················· ···················· ···················· ··A·················
M 7028 hCoV-19/England/ALDP-A2E48E/2020|EPI_ISL_586582|20 ···················· ···················· ···················· ···················· ···················· ····················
M 7029 hCoV-19/England/ALDP-A2DA50/2020|EPI_ISL_586583|20 ···················· ···················· ···················· ···················· ···················· ····················
M 7030 hCoV-19/England/ALDP-A2D3DD/2020|EPI_ISL_586584|20 ···················· ···················· ···················· ···················· ···················· ····················
M 7031 hCoV-19/England/ALDP-A2D66B/2020|EPI_ISL_586585|20 ···················· ···················· ···················· ···················· ···················· ····················
M 7032 hCoV-19/England/ALDP-A2E66A/2020|EPI_ISL_586586|20 ···················· ···················· ···················· ···················· ···················· ····················
M 7033 hCoV-19/England/ALDP-A2D416/2020|EPI_ISL_586588|20 ···················· ···················· ···················· ···················· ···················· ····················
M 7034 hCoV-19/England/ALDP-A2DDC0/2020|EPI_ISL_586589|20 ···················· ···················· ···················· ···················· ···················· ····················
M 7035 hCoV-19/England/ALDP-A2D452/2020|EPI_ISL_586590|20 ···················· ···················· ···················· ···················· ···················· ····················
M 7036 hCoV-19/England/ALDP-A2E284/2020|EPI_ISL_586591|20 ···················· ···················· ···················· ···················· ···················· ····················
M 7037 hCoV-19/England/ALDP-A2D5E6/2020|EPI_ISL_586592|20 ···················· ·A·················· ···················· ···················· ···················· ··A·················
M 7038 hCoV-19/England/ALDP-A2D777/2020|EPI_ISL_586593|20 ···················· ···················· ···················· ···················· ···················· ····················
M 7039 hCoV-19/England/ALDP-A2E2A2/2020|EPI_ISL_586595|20 ···················· ···················· ···················· ···················· ···················· ····················
M 7040 hCoV-19/England/ALDP-A2D522/2020|EPI_ISL_586596|20 ···················· ···················· ···················· ···················· ···················· ····················
M 7041 hCoV-19/England/ALDP-A2DE72/2020|EPI_ISL_586597|20 ···················· ···················· ···················· ···················· ···················· ····················
M 7042 hCoV-19/England/ALDP-A2DEAF/2020|EPI_ISL_586598|20 ···················· ···················· ···················· ···················· ···················· ····················
M 7043 hCoV-19/England/ALDP-A2DE45/2020|EPI_ISL_586599|20 ···················· ···················· ···················· ···················· ···················· ····················

```
M    7044 hCoV-19/England/ALDP-9FD656/2020|EPI_ISL_587801|20    ····················  ····················  ·····················  ····················  ····················  ····················
M    7045 hCoV-19/England/ALDP-9FCD34/2020|EPI_ISL_587804|20    ····················  ····················  ·····················  ····················  ····················  ····················
M    7046 hCoV-19/England/ALDP-9FD02B/2020|EPI_ISL_587805|20    ····················  ····················  ·····················  ····················  ····················  ····················
M    7047 hCoV-19/England/ALDP-9FD79F/2020|EPI_ISL_587806|20    ····················  ·A··················  ·····················  ····················  ····················  ··A·················
M    7048 hCoV-19/England/ALDP-9FCB0D/2020|EPI_ISL_587807|20    ····················  ····················  ·····················  ····················  ····················  ····················
M    7049 hCoV-19/England/ALDP-9FCB58/2020|EPI_ISL_587808|20    ····················  ····················  ·····················  ····················  ····················  ····················
M    7050 hCoV-19/England/ALDP-9FCC73/2020|EPI_ISL_587809|20    ····················  ····················  ·····················  ····················  ····················  ····················
M    7051 hCoV-19/England/ALDP-9FD850/2020|EPI_ISL_587810|20    ····················  ····················  ·····················  ····················  ····················  ····················
M    7052 hCoV-19/England/ALDP-9FCD07/2020|EPI_ISL_587811|20    ····················  ····················  ·····················  ····················  ····················  ····················
M    7053 hCoV-19/England/ALDP-9FD46B/2020|EPI_ISL_587813|20    ····················  ····················  ·····················  ····················  ····················  ····················
M    7054 hCoV-19/England/ALDP-9FD708/2020|EPI_ISL_587814|20    ····················  ····················  ·····················  ····················  ····················  ····················
M    7055 hCoV-19/England/ALDP-9FD38C/2020|EPI_ISL_587815|20    ····················  ····················  ·····················  ····················  ····················  ····················
M    7056 hCoV-19/England/ALDP-9FD182/2020|EPI_ISL_587817|20    ····················  ····················  ·····················  ····················  ····················  ····················
M    7057 hCoV-19/England/ALDP-9FD629/2020|EPI_ISL_587819|20    ····················  ····················  ·····················  ····················  ····················  ····················
M    7058 hCoV-19/England/ALDP-9FC842/2020|EPI_ISL_587820|20    ····················  ····················  ·····················  ····················  ····················  ····················
M    7059 hCoV-19/England/ALDP-9FD1CE/2020|EPI_ISL_587821|20    ····················  ····················  ·····················  ····················  ····················  ····················
M    7060 hCoV-19/England/ALDP-9FD4B6/2020|EPI_ISL_587822|20    ····················  ····················  ·····················  ····················  ····················  ····················
M    7061 hCoV-19/England/ALDP-9FDA96/2020|EPI_ISL_587825|20    ····················  ····················  ·····················  ····················  ····················  ····················
M    7062 hCoV-19/England/ALDP-9FDD06/2020|EPI_ISL_587826|20    ····················  ····················  ·····················  ····················  ····················  ····················
M    7063 hCoV-19/England/ALDP-9FCD8F/2020|EPI_ISL_587827|20    ····················  ····················  ·····················  ····················  ····················  ····················
M    7064 hCoV-19/England/ALDP-9FDE7C/2020|EPI_ISL_587828|20    ····················  ····················  ·····················  ····················  ····················  ····················
M    7065 hCoV-19/England/ALDP-9FC8F7/2020|EPI_ISL_587829|20    ····················  ····················  ·····················  ····················  ····················  ····················
M    7066 hCoV-19/England/ALDP-9FCF6B/2020|EPI_ISL_587830|20    ····················  ····················  ·····················  ····················  ····················  ····················
M    7067 hCoV-19/England/ALDP-9FDDF7/2020|EPI_ISL_587853|20    ····················  ····················  ·····················  ····················  ····················  ····················
M    7068 hCoV-19/England/ALDP-9FD9E4/2020|EPI_ISL_587854|20    ····················  ····················  ·····················  ····················  ····················  ····················
M    7069 hCoV-19/England/ALDP-9FD814/2020|EPI_ISL_587855|20    ····················  ····················  ·····················  ····················  ····················  ····················
M    7070 hCoV-19/England/ALDP-9FCA3D/2020|EPI_ISL_587856|20    ····················  ····················  ·····················  ····················  ····················  ····················
M    7071 hCoV-19/England/ALDP-9FD37D/2020|EPI_ISL_587857|20    ····················  ····················  ·····················  ····················  ····················  ····················
M    7072 hCoV-19/England/ALDP-9FDD9D/2020|EPI_ISL_587859|20    ····················  ····················  ·····················  ····················  ····················  ····················
M    7073 hCoV-19/England/ALDP-9FD595/2020|EPI_ISL_587860|20    ····················  ····················  ·····················  ····················  ····················  ····················
M    7074 hCoV-19/England/ALDP-9FCAD3/2020|EPI_ISL_587861|20    ····················  ····················  ·····················  ····················  ····················  ····················
M    7075 hCoV-19/England/ALDP-9FD6FC/2020|EPI_ISL_587862|20    ····················  ····················  ·····················  ····················  ····················  ····················
M    7076 hCoV-19/England/ALDP-9FCD61/2020|EPI_ISL_587863|20    ····················  ····················  ·····················  ····················  ····················  ····················
M    7077 hCoV-19/England/ALDP-9FC7CD/2020|EPI_ISL_587866|20    ····················  ····················  ·····················  ····················  ····················  ····················
M    7078 hCoV-19/England/ALDP-9FDEB8/2020|EPI_ISL_587875|20    ····················  ····················  ·····················  ····················  ····················  ····················
M    7079 hCoV-19/England/ALDP-9FCB85/2020|EPI_ISL_587876|20    ····················  ····················  ·····················  ····················  ····················  ····················
M    7080 hCoV-19/England/ALDP-9FD9C6/2020|EPI_ISL_587878|20    ····················  ····················  ·····················  ····················  ····················  ····················
M    7081 hCoV-19/England/ALDP-9FD920/2020|EPI_ISL_587879|20    ····················  ····················  ·····················  ····················  ····················  ····················
M    7082 hCoV-19/England/ALDP-9FD410/2020|EPI_ISL_587882|20    ····················  ····················  ·····················  ····················  ····················  ····················
M    7083 hCoV-19/England/ALDP-9FCF89/2020|EPI_ISL_587884|20    ····················  ····················  ·····················  ····················  ····················  ····················
M    7084 hCoV-19/England/ALDP-9FCDE9/2020|EPI_ISL_587885|20    ····················  ····················  ·····················  ····················  ······T·············  ····················
M    7085 hCoV-19/England/ALDP-9FC88E/2020|EPI_ISL_587886|20    ····················  ····················  ·····················  ····················  ····················  ····················
M    7086 hCoV-19/England/ALDP-9FCDF8/2020|EPI_ISL_587887|20    ····················  ····················  ·····················  ····················  ····················  ····················
M    7087 hCoV-19/England/ALDP-9FCA88/2020|EPI_ISL_587888|20    ····················  ····················  ·····················  ····················  ····················  ····················
M    7088 hCoV-19/England/ALDP-9FCCCE/2020|EPI_ISL_587890|20    ····················  ····················  ·····················  ····················  ····················  ····················
M    7089 hCoV-19/England/ALDP-9FDEE5/2020|EPI_ISL_587891|20    ····················  ····················  ·····················  ····················  ····················  ····················
M    7090 hCoV-19/England/ALDP-9FDCBE/2020|EPI_ISL_587892|20    ····················  ····················  ·····················  ····················  ····················  ····················
M    7091 hCoV-19/England/ALDP-9FD1EC/2020|EPI_ISL_587893|20    ····················  ····················  ·····················  ····················  ····················  ····················
M    7092 hCoV-19/England/ALDP-9FD304/2020|EPI_ISL_587894|20    ····················  ····················  ·····················  ····················  ····················  ····················
M    7093 hCoV-19/England/ALDP-9FDCEB/2020|EPI_ISL_587895|20    ····················  ····················  ·····················  ····················  ····················  ····················
M    7094 hCoV-19/England/ALDP-9FD225/2020|EPI_ISL_587896|20    ····················  ····················  ·····················  ····················  ····················  ····················
M    7095 hCoV-19/England/ALDP-9FDB48/2020|EPI_ISL_587897|20    ····················  ····················  ·····················  ····················  ····················  ····················
M    7096 hCoV-19/England/ALDP-9FCB67/2020|EPI_ISL_587898|20    ····················  ····················  ·····················  ····················  ····················  ····················
M    7097 hCoV-19/England/ALDP-9FDD24/2020|EPI_ISL_587899|20    ····················  ····················  ·····················  ····················  ····················  ····················
M    7098 hCoV-19/USA/MT-RML-7/2020|EPI_ISL_591054|2020-07-1     ····················  ·A··················  ·····················  ····················  ····················  ····················
M    7099 hCoV-19/USA/MT-RML-8/2020|EPI_ISL_591055|2020-07-1     ····················  ·A··················  ·····················  ····················  ····················  ····················
M    7100 hCoV-19/USA/MT-RML-9/2020|EPI_ISL_591056|2020-07-2     ····················  ·A··················  ·····················  ····················  ····················  ····················
M    7102 hCoV-19/DRC/10248/2020|EPI_ISL_591087|2020-05-22      ····················  ····················  ·····················  ····················  ····················  ····················
M    7104 hCoV-19/Australia/VIC14202/2020|EPI_ISL_592386|202    ····················  ····················  ·····················  ····················  ····················  ····················
M    7105 hCoV-19/Australia/VIC14203/2020|EPI_ISL_592387|202    ····················  ····················  ·····················  ····················  ····················  ····················
M    7106 hCoV-19/Australia/VIC14206/2020|EPI_ISL_592388|202    ····················  ····················  ·····················  ····················  ····················  ····················
M    7108 hCoV-19/Australia/VIC14209/2020|EPI_ISL_592390|202    ····················  ····················  ·····················  ····················  ····················  ····················
M    7109 hCoV-19/Australia/VIC14210/2020|EPI_ISL_592391|202    ····················  ····················  ·····················  ····················  ·····W··············  ····················
M    7110 hCoV-19/Australia/VIC14211/2020|EPI_ISL_592392|202    ····················  ····················  ·····················  ····················  ····················  ····················
M    7111 hCoV-19/Australia/VIC14213/2020|EPI_ISL_592393|202    ····················  ····················  ·····················  ····················  ····················  ····················
M    7112 hCoV-19/Australia/VIC14214/2020|EPI_ISL_592394|202    ····················  ····················  ·····················  ····················  ····················  ····················
M    7113 hCoV-19/Australia/VIC14217/2020|EPI_ISL_592395|202    ····················  ····················  ·····················  ····················  ····················  ····················
M    7114 hCoV-19/Australia/VIC14218/2020|EPI_ISL_592396|202    ····················  ····················  ·····················  ····················  ····················  ····················
M    7115 hCoV-19/Australia/VIC14219/2020|EPI_ISL_592397|202    ····················  ····················  ·····················  ····················  ····················  ····················
M    7116 hCoV-19/Australia/VIC14222/2020|EPI_ISL_592398|202    ····················  ····················  ·····················  ····················  ····················  ····················
M    7117 hCoV-19/Australia/VIC14223/2020|EPI_ISL_592399|202    ····················  ····················  ·····················  ····················  ····················  ····················
M    7118 hCoV-19/Australia/QLD1081/2020|EPI_ISL_593600|2020    ····················  ····················  ·····················  ····················  ····················  ····················
M    7119 hCoV-19/Australia/QLD1082/2020|EPI_ISL_593601|2020    ····················  ····················  ·····················  ····················  ····················  ····················
M    7120 hCoV-19/Australia/QLD1083/2020|EPI_ISL_593602|2020    ····················  ····················  ·····················  ····················  ····················  ····················
M    7121 hCoV-19/Australia/QLD1084/2020|EPI_ISL_593603|2020    ····················  ····················  ·····················  ····················  ····················  ····················
M    7122 hCoV-19/Australia/QLD1085/2020|EPI_ISL_593604|2020    ····················  ····················  ·····················  ····················  ····················  ····················
M    7123 hCoV-19/Australia/QLD1086/2020|EPI_ISL_593605|2020    ····················  ····················  ·····················  ····················  ····················  ····················
M    7124 hCoV-19/Australia/QLD1087/2020|EPI_ISL_593606|2020    ····················  ····················  ·····················  ····················  ····················  ····················
M    7125 hCoV-19/Australia/QLD1088/2020|EPI_ISL_593607|2020    ····················  ····················  ·····················  ····················  ····················  ····················
M    7126 hCoV-19/Australia/QLD1089/2020|EPI_ISL_593608|2020    ····················  ····················  ·····················  ····················  ····················  ····················
```

```
M	7127 hCoV-19/Australia/QLD1090/2020|EPI_ISL_593609|2020	····················	····················	·····················	····················	····················	····················
M	7128 hCoV-19/Australia/QLD1095/2020|EPI_ISL_593614|2020	····················	····················	·····················	····················	····················	····················
M	7129 hCoV-19/Australia/QLD1096/2020|EPI_ISL_593615|2020	····················	····················	·····················	····················	····················	····················
M	7130 hCoV-19/Australia/QLD1097/2020|EPI_ISL_593616|2020	····················	····················	·····················	····················	····················	····················
M	7131 hCoV-19/Australia/QLD1101/2020|EPI_ISL_593618|2020	····················	····················	·····················	····················	····················	····················
M	7132 hCoV-19/Australia/QLD1102/2020|EPI_ISL_593619|2020	····················	····················	·····················	····················	····················	····················
M	7133 hCoV-19/Australia/QLD1103/2020|EPI_ISL_593620|2020	····················	·A··················	·····················	····················	····················	····················
M	7134 hCoV-19/Australia/QLD1104/2020|EPI_ISL_593621|2020	····················	····················	·····················	····················	····················	····················
M	7135 hCoV-19/Australia/QLD1105/2020|EPI_ISL_593622|2020	····················	····················	·····················	····················	····················	····················
M	7136 hCoV-19/Australia/QLD1106/2020|EPI_ISL_593623|2020	····················	····················	·····················	····················	····················	····················
M	7137 hCoV-19/Australia/QLD1107/2020|EPI_ISL_593624|2020	····················	····················	·····················	····················	····················	····················
M	7138 hCoV-19/Australia/QLD1108/2020|EPI_ISL_593625|2020	····················	····················	·····················	····················	····················	····················
M	7139 hCoV-19/Australia/QLD1109/2020|EPI_ISL_593626|2020	····················	····················	·····················	····················	····················	····················
M	7140 hCoV-19/Australia/QLD1110/2020|EPI_ISL_593627|2020	····················	····················	·····················	····················	····················	····················
M	7141 hCoV-19/Australia/QLD1111/2020|EPI_ISL_593628|2020	····················	····················	·····················	····················	····················	····················
M	7142 hCoV-19/Australia/QLD1112/2020|EPI_ISL_593629|2020	····················	····················	·····················	····················	····················	····················
M	7143 hCoV-19/Wales/MILK-9EF4A8/2020|EPI_ISL_594920|2020	····················	····················	·····················	····················	····················	····················
M	7144 hCoV-19/Wales/MILK-9EF4C6/2020|EPI_ISL_594921|2020	····················	····················	·····················	····················	····················	····················
M	7145 hCoV-19/Wales/MILK-9EF4E4/2020|EPI_ISL_594922|2020	····················	····················	·····················	····················	····················	····················
M	7146 hCoV-19/England/MILK-9EF5F0/2020|EPI_ISL_594923|20	····················	····················	·····················	····················	····················	····················
M	7147 hCoV-19/England/MILK-9EF61B/2020|EPI_ISL_594924|20	····················	····················	·····················	····················	····················	····················
M	7148 hCoV-19/England/MILK-9EF639/2020|EPI_ISL_594926|20	····················	····················	·····················	····················	····················	····················
M	7149 hCoV-19/England/MILK-9EF666/2020|EPI_ISL_594927|20	····················	····················	·····················	····················	····················	····················
M	7150 hCoV-19/Wales/MILK-9EF675/2020|EPI_ISL_594928|2020	····················	····················	·····················	····················	····················	····················
M	7151 hCoV-19/England/MILK-9EF684/2020|EPI_ISL_594929|20	····················	····················	·····················	····················	····················	····················
M	7152 hCoV-19/England/MILK-9EF6A2/2020|EPI_ISL_594930|20	····················	·A··················	·····················	····················	····················	···A················
M	7153 hCoV-19/England/MILK-9EF6EE/2020|EPI_ISL_594931|20	····················	····················	·····················	····················	····················	····················
M	7154 hCoV-19/England/MILK-9EF6FD/2020|EPI_ISL_594932|20	····················	·A··················	·····················	····················	····················	····················
M	7155 hCoV-19/England/MILK-9EF709/2020|EPI_ISL_594933|20	····················	····················	·····················	····················	····················	····················
M	7156 hCoV-19/England/MILK-9EF718/2020|EPI_ISL_594934|20	····················	····················	·····················	····················	····················	····················
M	7157 hCoV-19/England/MILK-9EF772/2020|EPI_ISL_594937|20	····················	····················	·····················	····················	····················	····················
M	7158 hCoV-19/England/MILK-9EF781/2020|EPI_ISL_594938|20	····················	····················	·····················	····················	····················	····················
M	7159 hCoV-19/England/MILK-9EF790/2020|EPI_ISL_594939|20	····················	····················	·····················	····················	····················	····················
M	7160 hCoV-19/England/MILK-9EF7AF/2020|EPI_ISL_594940|20	····················	····················	·····················	····················	····················	····················
M	7161 hCoV-19/England/MILK-9EF806/2020|EPI_ISL_594945|20	····················	····················	·····················	····················	····················	····················
M	7162 hCoV-19/England/MILK-9EF815/2020|EPI_ISL_594946|20	····················	·A··················	·····················	····················	····················	···A················
M	7163 hCoV-19/England/MILK-9EF824/2020|EPI_ISL_594947|20	····················	····················	·····················	····················	····················	····················
M	7164 hCoV-19/England/MILK-9EF833/2020|EPI_ISL_594948|20	····················	·A··················	·····················	····················	····················	···A················
M	7165 hCoV-19/England/MILK-9EF8AC/2020|EPI_ISL_594953|20	····················	····················	·····················	····················	······T·············	····················
M	7166 hCoV-19/England/MILK-9EF8BB/2020|EPI_ISL_594954|20	····················	····················	·····················	····················	····················	····················
M	7167 hCoV-19/England/MILK-9EF8E8/2020|EPI_ISL_594955|20	····················	····················	·····················	····················	····················	····················
M	7168 hCoV-19/England/MILK-9EF8F7/2020|EPI_ISL_594956|20	····················	····················	·····················	····················	····················	····················
M	7169 hCoV-19/England/LIVE-99BFC/2020|EPI_ISL_584607|202	····················	····················	·····················	····················	····················	····················
M	7170 hCoV-19/England/LIVE-99C80/2020|EPI_ISL_584608|202	····················	····················	·····················	····················	····················	····················
M	7171 hCoV-19/England/LIVE-99E20/2020|EPI_ISL_584611|202	····················	····················	·····················	····················	····················	····················
M	7172 hCoV-19/England/LIVE-99E6C/2020|EPI_ISL_584612|202	····················	····················	·····················	····················	····················	····················
M	7173 hCoV-19/England/LIVE-99E7B/2020|EPI_ISL_584613|202	····················	····················	·····················	····················	····················	····················
M	7175 hCoV-19/England/LIVE-A3F20/2020|EPI_ISL_584617|202	····················	····················	·····················	····················	····················	····················
M	7176 hCoV-19/England/LIVE-A404A/2020|EPI_ISL_584618|202	····················	····················	·····················	····················	····················	····················
M	7179 hCoV-19/England/LOND-124D8DD/2020|EPI_ISL_584622|2	····················	····················	·····················	····················	····················	····················
M	7180 hCoV-19/England/LOND-124D934/2020|EPI_ISL_584624|2	····················	····················	·····················	····················	····················	····················
M	7181 hCoV-19/England/LOND-124D952/2020|EPI_ISL_584625|2	····················	····················	·····················	····················	····················	····················
M	7182 hCoV-19/England/LOND-124D98F/2020|EPI_ISL_584627|2	····················	····················	·····················	····················	····················	····················
M	7183 hCoV-19/England/LOND-124D9AD/2020|EPI_ISL_584628|2	····················	····················	·····················	····················	····················	····················
M	7185 hCoV-19/England/LOND-124DA13/2020|EPI_ISL_584631|2	····················	····················	·····················	····················	····················	····················
M	7186 hCoV-19/England/LOND-124DA7D/2020|EPI_ISL_584636|2	····················	····················	·····················	····················	····················	····················
M	7187 hCoV-19/England/LOND-124DAF5/2020|EPI_ISL_584640|2	····················	····················	·····················	····················	····················	····················
M	7188 hCoV-19/Northern Ireland/NIRE-10878A/2020|EPI_ISL_	····················	····················	·····················	····················	····················	····················
M	7189 hCoV-19/Northern Ireland/NIRE-1087B7/2020|EPI_ISL_	····················	····················	·····················	····················	····················	····················
M	7190 hCoV-19/Northern Ireland/NIRE-10881E/2020|EPI_ISL_	····················	····················	·····················	····················	····················	····················
M	7191 hCoV-19/Northern Ireland/NIRE-10883C/2020|EPI_ISL_	····················	····················	·····················	····················	····················	····················
M	7192 hCoV-19/Northern Ireland/NIRE-10885A/2020|EPI_ISL_	····················	····················	·····················	····················	N···················	····················
M	7193 hCoV-19/Northern Ireland/NIRE-108869/2020|EPI_ISL_	····················	····················	·····················	····················	····················	····················
M	7194 hCoV-19/Northern Ireland/NIRE-108887/2020|EPI_ISL_	····················	····················	·····················	····················	····················	····················
M	7195 hCoV-19/Northern Ireland/NIRE-108896/2020|EPI_ISL_	····················	····················	·····················	····················	····················	····················
M	7196 hCoV-19/Northern Ireland/NIRE-1088A5/2020|EPI_ISL_	····················	····················	·····················	····················	····················	····················
M	7197 hCoV-19/Wales/PHWC-16F01D/2020|EPI_ISL_585900|2020	····················	····················	·····················	····················	····················	····················
M	7198 hCoV-19/Wales/PHWC-16F03B/2020|EPI_ISL_585902|2020	····················	····················	·····················	····················	····················	····················
M	7199 hCoV-19/Wales/PHWC-16F04A/2020|EPI_ISL_585903|2020	····················	····················	·····················	····················	····················	····················
M	7200 hCoV-19/Wales/PHWC-16F059/2020|EPI_ISL_585904|2020	····················	····················	·····················	····················	····················	····················
M	7201 hCoV-19/Wales/PHWC-16F068/2020|EPI_ISL_585905|2020	····················	····················	·····················	····················	····················	····················
M	7202 hCoV-19/Wales/PHWC-16F077/2020|EPI_ISL_585906|2020	····················	····················	·····················	····················	····················	····················
M	7203 hCoV-19/Wales/PHWC-16F095/2020|EPI_ISL_585908|2020	····················	····················	·····················	····················	····················	····················
M	7204 hCoV-19/Wales/PHWC-16F0A4/2020|EPI_ISL_585909|2020	····················	····················	·····················	····················	····················	····················
M	7205 hCoV-19/Wales/PHWC-16F0C2/2020|EPI_ISL_585911|2020	····················	····················	·····················	····················	····················	····················
M	7206 hCoV-19/Wales/PHWC-16F0D1/2020|EPI_ISL_585912|2020	····················	····················	·····················	····················	····················	····················
M	7207 hCoV-19/Wales/PHWC-16F0E0/2020|EPI_ISL_585913|2020	····················	····················	·····················	····················	····················	····················
M	7208 hCoV-19/Wales/PHWC-16F0FF/2020|EPI_ISL_585914|2020	····················	····················	·····················	····················	····················	····················
M	7209 hCoV-19/Wales/PHWC-16F10B/2020|EPI_ISL_585915|2020	····················	····················	·····················	····················	····················	····················
M	7210 hCoV-19/Wales/PHWC-16F129/2020|EPI_ISL_585916|2020	····················	····················	·····················	····················	····················	····················
```

| | | | | | | | |
|---|---|---|---|---|---|---|---|
| M | 7211 hCoV-19/Wales/PHWC-16F138/2020\|EPI_ISL_585917\|2020 | ···················· | ···················· | ···················· | ···················· | ···················· | ···················· |
| M | 7212 hCoV-19/Wales/PHWC-16F147/2020\|EPI_ISL_585918\|2020 | ···················· | ···················· | ···················· | ···················· | ···················· | ···················· |
| M | 7213 hCoV-19/Wales/PHWC-16F156/2020\|EPI_ISL_585919\|2020 | ···················· | ···················· | ···················· | ···················· | ···················· | ···················· |
| M | 7214 hCoV-19/Wales/PHWC-16F165/2020\|EPI_ISL_585920\|2020 | ···················· | ···················· | ···················· | ···················· | ···················· | ···················· |
| M | 7215 hCoV-19/Wales/PHWC-16F174/2020\|EPI_ISL_585921\|2020 | ···················· | ···················· | ···················· | ···················· | ···················· | ···················· |
| M | 7216 hCoV-19/Wales/PHWC-16F183/2020\|EPI_ISL_585922\|2020 | ···················· | ···················· | ···················· | ···················· | ···················· | ···················· |
| M | 7217 hCoV-19/Wales/PHWC-16F192/2020\|EPI_ISL_585923\|2020 | ···················· | ···················· | ···················· | ···················· | ···················· | ···················· |
| M | 7218 hCoV-19/Wales/PHWC-16F1A1/2020\|EPI_ISL_585924\|2020 | ···················· | ···················· | ···················· | ···················· | ···················· | ···················· |
| M | 7219 hCoV-19/Wales/PHWC-16F1CF/2020\|EPI_ISL_585926\|2020 | ···················· | ···················· | ···················· | ···················· | ···················· | ···················· |
| M | 7220 hCoV-19/Wales/PHWC-16F1DE/2020\|EPI_ISL_585927\|2020 | ···················· | ···················· | ···················· | ···················· | ···················· | ···················· |
| M | 7221 hCoV-19/Wales/PHWC-16F1ED/2020\|EPI_ISL_585928\|2020 | ···················· | ···················· | ···················· | ···················· | ···················· | ···················· |
| M | 7222 hCoV-19/Wales/PHWC-16F1FC/2020\|EPI_ISL_585929\|2020 | ···················· | ···················· | ···················· | ···················· | ···················· | ···················· |
| M | 7223 hCoV-19/Wales/PHWC-16F208/2020\|EPI_ISL_585930\|2020 | ···················· | ···················· | ···················· | ···················· | ···················· | ···················· |
| M | 7224 hCoV-19/Wales/PHWC-16F217/2020\|EPI_ISL_585931\|2020 | ···················· | ···················· | ···················· | ···················· | ···················· | ···················· |
| M | 7225 hCoV-19/Wales/PHWC-16F226/2020\|EPI_ISL_585932\|2020 | ···················· | ···················· | ···················· | ···················· | ···················· | ···················· |
| M | 7226 hCoV-19/Wales/PHWC-16F235/2020\|EPI_ISL_585933\|2020 | ···················· | ···················· | ···················· | ···················· | ···················· | ···················· |
| M | 7227 hCoV-19/Wales/PHWC-16F244/2020\|EPI_ISL_585934\|2020 | ···················· | ···················· | ···················· | ···················· | ···················· | ···················· |
| M | 7228 hCoV-19/Wales/PHWC-16F253/2020\|EPI_ISL_585935\|2020 | ···················· | ···················· | ···················· | ···················· | ···················· | ···················· |
| M | 7229 hCoV-19/Wales/PHWC-16F262/2020\|EPI_ISL_585936\|2020 | ···················· | ···················· | ···················· | ···················· | ···················· | ···················· |
| M | 7230 hCoV-19/Wales/PHWC-16F271/2020\|EPI_ISL_585937\|2020 | ···················· | ···················· | ···················· | ···················· | ···················· | ···················· |
| M | 7231 hCoV-19/Wales/PHWC-16F280/2020\|EPI_ISL_585938\|2020 | ···················· | ···················· | ···················· | ···················· | ···················· | ···················· |
| M | 7232 hCoV-19/Wales/PHWC-16F29F/2020\|EPI_ISL_585939\|2020 | ···················· | ···················· | ···················· | ···················· | ···················· | ···················· |
| M | 7233 hCoV-19/Wales/PHWC-16F2AE/2020\|EPI_ISL_585940\|2020 | ···················· | ···················· | ···················· | ···················· | ···················· | ···················· |
| M | 7234 hCoV-19/Wales/PHWC-16F2BD/2020\|EPI_ISL_585941\|2020 | ···················· | ···················· | ···················· | ···················· | ···················· | ···················· |
| M | 7235 hCoV-19/Wales/PHWC-16F2CC/2020\|EPI_ISL_585942\|2020 | ···················· | ···················· | ···················· | ···················· | ···················· | ···················· |
| M | 7236 hCoV-19/Wales/PHWC-16F2DB/2020\|EPI_ISL_585943\|2020 | ···················· | ···················· | ···················· | ···················· | ···················· | ···················· |
| M | 7237 hCoV-19/Wales/PHWC-16F2EA/2020\|EPI_ISL_585944\|2020 | ···················· | ···················· | ···················· | ···················· | ···················· | ···················· |
| M | 7238 hCoV-19/Wales/PHWC-16F2F9/2020\|EPI_ISL_585945\|2020 | ···················· | ···················· | ···················· | ···················· | ···················· | ···················· |
| M | 7239 hCoV-19/Wales/PHWC-16F305/2020\|EPI_ISL_585946\|2020 | ···················· | ···················· | ···················· | ···················· | ···················· | ···················· |
| M | 7240 hCoV-19/Wales/PHWC-16F314/2020\|EPI_ISL_585947\|2020 | ···················· | ···················· | ···················· | ···················· | ···················· | ···················· |
| M | 7241 hCoV-19/Wales/PHWC-16F323/2020\|EPI_ISL_585948\|2020 | ···················· | ···················· | ···················· | ···················· | ···················· | ···················· |
| M | 7242 hCoV-19/Wales/PHWC-16F332/2020\|EPI_ISL_585949\|2020 | ···················· | ···················· | ···················· | ···················· | ···················· | ···················· |
| M | 7243 hCoV-19/Wales/PHWC-16F341/2020\|EPI_ISL_585950\|2020 | ···················· | ···················· | ···················· | ···················· | ···················· | ···················· |
| M | 7245 hCoV-19/Wales/PHWC-16F37E/2020\|EPI_ISL_585953\|2020 | ···················· | ···················· | ···················· | ···················· | ···················· | ···················· |
| M | 7246 hCoV-19/Wales/PHWC-16F38D/2020\|EPI_ISL_585954\|2020 | ···················· | ···················· | ···················· | ···················· | ···················· | ···················· |
| M | 7247 hCoV-19/Wales/PHWC-16F39C/2020\|EPI_ISL_585955\|2020 | ···················· | ···················· | ···················· | ···················· | ···················· | ···················· |
| M | 7248 hCoV-19/Wales/PHWC-16F3AB/2020\|EPI_ISL_585956\|2020 | ···················· | ···················· | ···················· | ···················· | ···················· | ···················· |
| M | 7249 hCoV-19/Wales/PHWC-16F3BA/2020\|EPI_ISL_585957\|2020 | ···················· | ···················· | ···················· | ···················· | ···················· | ···················· |
| M | 7250 hCoV-19/Wales/PHWC-16F3C9/2020\|EPI_ISL_585958\|2020 | ···················· | ···················· | ···················· | ···················· | ···················· | ···················· |
| M | 7251 hCoV-19/Wales/PHWC-16F3D8/2020\|EPI_ISL_585959\|2020 | ···················· | ···················· | ···················· | ···················· | ···················· | ···················· |
| M | 7252 hCoV-19/Wales/PHWC-16F3E7/2020\|EPI_ISL_585960\|2020 | ···················· | ···················· | ···················· | ···················· | ···················· | ···················· |
| M | 7253 hCoV-19/Wales/PHWC-16F3F6/2020\|EPI_ISL_585961\|2020 | ···················· | ···················· | ···················· | ···················· | ···················· | ···················· |
| M | 7254 hCoV-19/Wales/PHWC-16F402/2020\|EPI_ISL_585962\|2020 | ···················· | ···················· | ···················· | ···················· | ···················· | ···················· |
| M | 7255 hCoV-19/Wales/PHWC-16F411/2020\|EPI_ISL_585963\|2020 | ···················· | ···················· | ···················· | ···················· | ···················· | ···················· |
| M | 7256 hCoV-19/Wales/PHWC-16F420/2020\|EPI_ISL_585964\|2020 | ···················· | ···················· | ···················· | ···················· | ···················· | ···················· |
| M | 7257 hCoV-19/Wales/PHWC-16F43F/2020\|EPI_ISL_585965\|2020 | ···················· | ···················· | ···················· | ···················· | ···················· | ···················· |
| M | 7258 hCoV-19/Wales/PHWC-16F44E/2020\|EPI_ISL_585966\|2020 | ···················· | ···················· | ···················· | ···················· | ···················· | ···················· |
| M | 7259 hCoV-19/Wales/PHWC-16F45D/2020\|EPI_ISL_585967\|2020 | ···················· | ···················· | ···················· | ···················· | ···················· | ···················· |
| M | 7260 hCoV-19/Wales/PHWC-16F46C/2020\|EPI_ISL_585968\|2020 | ···················· | ···················· | ···················· | ···················· | ···················· | ···················· |
| M | 7261 hCoV-19/Wales/PHWC-16F48A/2020\|EPI_ISL_585970\|2020 | ···················· | ···················· | ···················· | ···················· | ···················· | ···················· |
| M | 7262 hCoV-19/Wales/PHWC-16F499/2020\|EPI_ISL_585971\|2020 | ···················· | ···················· | ···················· | ···················· | ···················· | ···················· |
| M | 7263 hCoV-19/Canada/ON-S947/2020\|EPI_ISL_586500\|2020-04 | ···················· | ···················· | ···················· | ···················· | ···················· | ···················· |
| M | 7264 hCoV-19/Canada/ON-S948/2020\|EPI_ISL_586501\|2020-04 | ···················· | ···················· | ···················· | ···················· | ···················· | ···················· |
| M | 7265 hCoV-19/Canada/ON-S949/2020\|EPI_ISL_586502\|2020-04 | ···················· | ···················· | ···················· | ···················· | ···················· | ···················· |
| M | 7266 hCoV-19/Canada/ON-S952/2020\|EPI_ISL_586504\|2020-04 | ···················· | ···················· | ···················· | ···················· | ···················· | ···················· |
| M | 7267 hCoV-19/Canada/ON-S96/2020\|EPI_ISL_586505\|2020-03- | ···················· | ···················· | ···················· | ···················· | ···················· | ···················· |
| M | 7268 hCoV-19/India/GJ-GBRC-398a/2020\|EPI_ISL_586513\|202 | ···················· | ·A·················· | ···················· | ···················· | ···················· | ··A················· |
| M | 7269 hCoV-19/India/GJ-GBRC-398b/2020\|EPI_ISL_586514\|202 | ···················· | ·A·················· | ···················· | ···················· | ···················· | ··A················· |
| M | 7270 hCoV-19/India/GJ-GBRC-399a/2020\|EPI_ISL_586515\|202 | ···················· | ·A·················· | ···················· | ···················· | ···················· | ··A················· |
| M | 7271 hCoV-19/India/GJ-GBRC-399b/2020\|EPI_ISL_586516\|202 | ···················· | ·A·················· | ···················· | ···················· | ···················· | ···················· |
| M | 7272 hCoV-19/India/GJ-GBRC-400a/2020\|EPI_ISL_586517\|202 | ···················· | ·A·················· | ···················· | ···················· | ···················· | ··A················· |
| M | 7273 hCoV-19/India/GJ-GBRC-400b/2020\|EPI_ISL_586518\|202 | ···················· | ···················· | ···················· | ···················· | ···················· | ···················· |
| M | 7274 hCoV-19/India/GJ-GBRC-401/2020\|EPI_ISL_586519\|2020 | ···················· | ···················· | ···················· | ···················· | ···················· | ···················· |
| M | 7275 hCoV-19/India/GJ-GBRC-402a/2020\|EPI_ISL_586520\|202 | ···················· | ·A·················· | ···················· | ···················· | ···················· | ··A················· |
| M | 7276 hCoV-19/India/GJ-GBRC-402b/2020\|EPI_ISL_586521\|202 | ···················· | ·A·················· | ···················· | ···················· | ···················· | ···················· |
| M | 7277 hCoV-19/India/GJ-GBRC-409a/2020\|EPI_ISL_586533\|202 | ···················· | ···················· | ···················· | ···················· | ···················· | ···················· |
| M | 7278 hCoV-19/India/GJ-GBRC-410/2020\|EPI_ISL_586535\|2020 | ···················· | ···················· | ···················· | ···················· | ···················· | ···················· |
| M | 7279 hCoV-19/India/GJ-GBRC-411/2020\|EPI_ISL_586536\|2020 | ···················· | ·A·················· | ···················· | ···················· | ···················· | ··A················· |
| M | 7280 hCoV-19/India/GJ-GBRC-412a/2020\|EPI_ISL_586537\|202 | ···················· | ···················· | ···················· | ···················· | ···················· | ···················· |
| M | 7281 hCoV-19/India/GJ-GBRC-412b/2020\|EPI_ISL_586538\|202 | ···················· | ·A·················· | ···················· | ···················· | ···················· | ···················· |
| M | 7282 hCoV-19/India/GJ-GBRC-413a/2020\|EPI_ISL_586539\|202 | ···················· | ·A·················· | ···················· | ···················· | ···················· | ··A················· |
| M | 7283 hCoV-19/India/GJ-GBRC-413b/2020\|EPI_ISL_586540\|202 | ···················· | ·A·················· | ···················· | ···················· | ···················· | ··A················· |
| M | 7284 hCoV-19/India/GJ-GBRC-414/2020\|EPI_ISL_586541\|2020 | ···················· | ···················· | ···················· | ···················· | ···················· | ···················· |
| M | 7285 hCoV-19/India/GJ-GBRC-415a/2020\|EPI_ISL_586542\|202 | ···················· | ···················· | ···················· | ···················· | ···················· | ···················· |
| M | 7286 hCoV-19/India/GJ-GBRC-415b/2020\|EPI_ISL_586543\|202 | ···················· | ···················· | ···················· | ···················· | ···················· | ···················· |
| M | 7287 hCoV-19/England/ALDP-9FD0C1/2020\|EPI_ISL_587831\|20 | ···················· | ···················· | ···················· | ···················· | ···················· | ···················· |
| M | 7288 hCoV-19/England/ALDP-9FD44D/2020\|EPI_ISL_587832\|20 | ···················· | ···················· | ···················· | ···················· | ···················· | ···················· |
| M | 7289 hCoV-19/England/ALDP-9FD674/2020\|EPI_ISL_587833\|20 | ···················· | ···················· | ···················· | ···················· | ···················· | ···················· |
| M | 7290 hCoV-19/England/ALDP-9FD717/2020\|EPI_ISL_587834\|20 | ···················· | ···················· | ···················· | ···················· | ···················· | ···················· |
| M | 7291 hCoV-19/England/ALDP-9FDE21/2020\|EPI_ISL_587835\|20 | ···················· | ···················· | ···················· | ···················· | ···················· | ···················· |

M 7292 hCoV-19/England/ALDP-9FCFF2/2020|EPI_ISL_587836|20 ···················· ···················· ···················· ···················· ···················· ····················
M 7293 hCoV-19/England/ALDP-9FD6B0/2020|EPI_ISL_587837|20 ···················· ···················· ···················· ···················· ···················· ····················
M 7294 hCoV-19/England/ALDP-9FD498/2020|EPI_ISL_587838|20 ···················· ···················· ···················· ···················· ···················· ····················
M 7295 hCoV-19/England/ALDP-9FC8CA/2020|EPI_ISL_587840|20 ···················· ···················· ···················· ···················· ···················· ····················
M 7296 hCoV-19/England/ALDP-9FD0A3/2020|EPI_ISL_587841|20 ···················· ···················· ···················· ···················· ···················· ····················
M 7297 hCoV-19/England/ALDP-9FD53B/2020|EPI_ISL_587843|20 ···················· ···················· ···················· ···················· ···················· ····················
M 7298 hCoV-19/England/ALDP-9FDD51/2020|EPI_ISL_587844|20 ···················· ···················· ···················· ···················· ···················· ····················
M 7299 hCoV-19/England/ALDP-9FD9B7/2020|EPI_ISL_587845|20 ···················· ···················· ···················· ···················· ···················· ····················
M 7300 hCoV-19/England/ALDP-9FC860/2020|EPI_ISL_587846|20 ···················· ···················· ···················· ···················· ···················· ····················
M 7301 hCoV-19/England/ALDP-9FC7AF/2020|EPI_ISL_587847|20 ···················· ···················· ···················· ···················· ···················· ····················
M 7302 hCoV-19/England/ALDP-9FDBFD/2020|EPI_ISL_587848|20 ···················· ···················· ···················· ···················· ···················· ····················
M 7303 hCoV-19/England/ALDP-9FDF5B/2020|EPI_ISL_587849|20 ···················· ···················· ···················· ···················· ···················· ····················
M 7304 hCoV-19/England/ALDP-9FD340/2020|EPI_ISL_587851|20 ···················· ···················· ···················· ···················· ···················· ····················
M 7305 hCoV-19/England/ALDP-9FCDDA/2020|EPI_ISL_587852|20 ···················· ···················· ···················· ···················· ···················· ····················
M 7306 hCoV-19/England/ALDP-9FDB2A/2020|EPI_ISL_587864|20 ···················· ···················· ···················· ···················· ···················· ····················
M 7307 hCoV-19/England/ALDP-9FDB1B/2020|EPI_ISL_587865|20 ···················· ···················· ···················· ···················· ···················· ····················
M 7308 hCoV-19/England/ALDP-9FCA5B/2020|EPI_ISL_587867|20 ···················· ···················· ···················· ···················· ···················· ····················
M 7309 hCoV-19/England/ALDP-9FCCDD/2020|EPI_ISL_587868|20 ···················· ···················· ···················· ···················· ···················· ····················
M 7310 hCoV-19/England/ALDP-9FDED6/2020|EPI_ISL_587869|20 ···················· ···················· ···················· ···················· ···················· ····················
M 7311 hCoV-19/England/ALDP-9FCB3A/2020|EPI_ISL_587870|20 ···················· ···················· ···················· ···················· ···················· ····················
M 7312 hCoV-19/England/ALDP-9FDC18/2020|EPI_ISL_587871|20 ···················· ···················· ···················· ···················· ···················· ····················
M 7313 hCoV-19/England/ALDP-9FD86F/2020|EPI_ISL_587872|20 ···················· ···················· ···················· ···················· ···················· ····················
M 7314 hCoV-19/England/ALDP-9FD568/2020|EPI_ISL_587873|20 ···················· ···················· ···················· ···················· ···················· ····················
M 7315 hCoV-19/England/ALDP-9FDE4F/2020|EPI_ISL_587874|20 ···················· ···················· ···················· ···················· ···················· ····················
M 7316 hCoV-19/England/QEUH-9FA695/2020|EPI_ISL_588106|20 ···················· ···················· ···················· ···················· ···················· ····················
M 7317 hCoV-19/England/QEUH-9FB3D9/2020|EPI_ISL_588107|20 ···················· ···················· ···················· ···················· ···················· ····················
M 7318 hCoV-19/England/QEUH-9FB096/2020|EPI_ISL_588108|20 ···················· ···················· ···················· ···················· ···················· ····················
M 7319 hCoV-19/England/QEUH-9FAAB7/2020|EPI_ISL_588109|20 ···················· ···················· ···················· ···················· ···················· ····················
M 7320 hCoV-19/England/QEUH-9FAB3C/2020|EPI_ISL_588110|20 ···················· ···················· ···················· ···················· ···················· ····················
M 7321 hCoV-19/England/QEUH-9FB315/2020|EPI_ISL_588111|20 ···················· ···················· ···················· ···················· ···T················ ····················
M 7322 hCoV-19/England/QEUH-9FA6C2/2020|EPI_ISL_588113|20 ···················· ···················· ···················· ···················· ···················· ····················
M 7323 hCoV-19/England/QEUH-9FA3DA/2020|EPI_ISL_588115|20 ···················· ···················· ···················· ···················· ···················· ····················
M 7324 hCoV-19/England/QEUH-9FACDF/2020|EPI_ISL_588116|20 ···················· ···················· ···················· ···················· ···················· ····················
M 7325 hCoV-19/England/QEUH-9FA8BD/2020|EPI_ISL_588117|20 ···················· ···················· ···················· ···················· ···T················ ····················
M 7326 hCoV-19/England/QEUH-9FB281/2020|EPI_ISL_588119|20 ···················· ···················· ···················· ···················· ···················· ····················
M 7327 hCoV-19/England/QEUH-9FAD09/2020|EPI_ISL_588120|20 ···················· ···················· ···················· ···················· ···················· ····················
M 7328 hCoV-19/England/QEUH-9FB209/2020|EPI_ISL_588121|20 ···················· ···················· ···················· ···················· ···················· ····················
M 7329 hCoV-19/England/QEUH-9FB342/2020|EPI_ISL_588122|20 ···················· ···················· ···················· ···················· ···················· ····················
M 7330 hCoV-19/England/QEUH-9FAA8A/2020|EPI_ISL_588123|20 ···················· ···················· ···················· ···················· ···················· ····················
M 7331 hCoV-19/England/QEUH-9FAB4B/2020|EPI_ISL_588124|20 ···················· ···················· ···················· ···················· ···················· ····················
M 7332 hCoV-19/England/QEUH-9FAC39/2020|EPI_ISL_588126|20 ···················· ·A·················· ···················· ···················· ···················· ··A·················
M 7333 hCoV-19/England/QEUH-9FB1C0/2020|EPI_ISL_588127|20 ···················· ···················· ···················· ···················· ···················· ····················
M 7334 hCoV-19/England/QEUH-9FA835/2020|EPI_ISL_588170|20 ···················· ···················· ···················· ···················· ···················· ····················
M 7335 hCoV-19/England/QEUH-9FAA6C/2020|EPI_ISL_588171|20 ···················· ·A·················· ···················· ···················· ···················· ····················
M 7336 hCoV-19/England/QEUH-9FA316/2020|EPI_ISL_588172|20 ···················· ···················· ···················· ···················· ···················· ····················
M 7337 hCoV-19/England/QEUH-9FA89F/2020|EPI_ISL_588173|20 ···················· ···················· ···················· ···················· ···················· ····················
M 7338 hCoV-19/England/QEUH-9FA6FF/2020|EPI_ISL_588174|20 ···················· ···················· ···················· ···················· ···················· ····················
M 7339 hCoV-19/England/QEUH-9FAC84/2020|EPI_ISL_588176|20 ···················· ···················· ···················· ···················· ···················· ····················
M 7340 hCoV-19/England/QEUH-9FB55B/2020|EPI_ISL_588177|20 ···················· ···················· ···················· ···················· ·····T·············· ····················
M 7341 hCoV-19/England/QEUH-9FA5D4/2020|EPI_ISL_588178|20 ···················· ·A·················· ···················· ···················· ···················· ····················
M 7342 hCoV-19/England/QEUH-9FAD54/2020|EPI_ISL_588179|20 ···················· ···················· ···················· ···················· ···················· ····················
M 7343 hCoV-19/England/QEUH-9FAF21/2020|EPI_ISL_588180|20 ···················· ···················· ···················· ···················· ···················· ····················
M 7344 hCoV-19/England/QEUH-9FAC57/2020|EPI_ISL_588181|20 ···················· ···················· ···················· ···················· ···················· ····················
M 7345 hCoV-19/England/QEUH-9FA7DE/2020|EPI_ISL_588182|20 ···················· ···················· ···················· ···················· ···················· ····················
M 7346 hCoV-19/England/QEUH-9FAEBB/2020|EPI_ISL_588183|20 ···················· ···················· ···················· ···················· ···················· ····················
M 7347 hCoV-19/England/QEUH-9FB2CD/2020|EPI_ISL_588184|20 ···················· ···················· ···················· ···················· ···················· ····················
M 7348 hCoV-19/England/QEUH-9FAC66/2020|EPI_ISL_588185|20 ···················· ···················· ···················· ···················· ···················· ····················
M 7349 hCoV-19/England/QEUH-9FA39E/2020|EPI_ISL_588186|20 ···················· ··G················· ···················· ···················· ···················· ····················
M 7350 hCoV-19/England/QEUH-9FA61D/2020|EPI_ISL_588187|20 ···················· ···················· ···················· ···················· ···················· ····················
M 7351 hCoV-19/England/QEUH-9FA62C/2020|EPI_ISL_588188|20 ···················· ···················· ···················· ···················· ···················· ····················
M 7352 hCoV-19/England/QEUH-9FADFA/2020|EPI_ISL_588189|20 ···················· ···················· ···················· ···················· ···················· ····················
M 7353 hCoV-19/England/QEUH-9FADAF/2020|EPI_ISL_588190|20 ···················· ···················· ···················· ···················· ···················· ····················
M 7354 hCoV-19/England/QEUH-9FA361/2020|EPI_ISL_588191|20 ···················· ···················· ···················· ···················· ···················· ····················
M 7355 hCoV-19/England/QEUH-9FB01E/2020|EPI_ISL_588192|20 ···················· ···················· ···················· ···················· ···················· ····················
M 7356 hCoV-19/England/QEUH-9FAA11/2020|EPI_ISL_588193|20 ···················· ···················· ···················· ···················· ···················· ····················
M 7357 hCoV-19/England/QEUH-9FAAF3/2020|EPI_ISL_588194|20 ···················· ···················· ···················· ···················· ···················· ····················
M 7358 hCoV-19/England/QEUH-9FAFB8/2020|EPI_ISL_588195|20 ···················· ···················· ···················· ···················· ···················· ····················
M 7359 hCoV-19/England/QEUH-9FAFC7/2020|EPI_ISL_588196|20 ···················· ···················· ···················· ···················· ···················· ····················
M 7360 hCoV-19/England/QEUH-9FB069/2020|EPI_ISL_588197|20 ···················· ···················· ···················· ···················· ···················· ····················
M 7361 hCoV-19/England/QEUH-9FAC93/2020|EPI_ISL_588199|20 ···················· ·A·················· ···················· ···················· ···················· ··A·················
M 7362 hCoV-19/England/ALDP-9EE3BB/2020|EPI_ISL_589415|20 ···················· ···················· ···················· ···················· ···················· ····················
M 7363 hCoV-19/England/ALDP-9E5B9E/2020|EPI_ISL_589416|20 ···················· ·A·················· ···················· ···················· ···················· ··A·················
M 7364 hCoV-19/England/MILK-9ECC67/2020|EPI_ISL_589417|20 ···················· ···················· ···················· ···················· ···················· ····················
M 7365 hCoV-19/England/ALDP-9ED4E6/2020|EPI_ISL_589418|20 ···················· ·A·················· ···················· ···················· ···················· ····················
M 7366 hCoV-19/England/ALDP-9EE2AF/2020|EPI_ISL_589419|20 ···················· ···················· ···················· ···················· ···················· ····················
M 7367 hCoV-19/England/ALDP-9E5897/2020|EPI_ISL_589420|20 ···················· ···················· ···················· ···················· ···················· ····················
M 7368 hCoV-19/England/ALDP-9ED246/2020|EPI_ISL_589421|20 ···················· ···················· ···················· ···················· ···················· ····················
M 7369 hCoV-19/England/ALDP-9E5CF5/2020|EPI_ISL_589437|20 ···················· ···················· ···················· ···················· ···················· ····················
M 7370 hCoV-19/England/ALDP-9ED61D/2020|EPI_ISL_589438|20 ···················· ···················· ···················· ···················· ···················· ····················
M 7371 hCoV-19/England/ALDP-9ECE34/2020|EPI_ISL_589439|20 ···················· ···················· ···················· ···················· ···················· ····················

| | | | | | | | | |
|---|---|---|---|---|---|---|---|---|
| M | 7372 | hCoV-19/England/MILK-9ECBC4/2020\|EPI_ISL_589440\|20 | .................... | .................... | ..................... | .................... | .................... | .................... |
| M | 7373 | hCoV-19/England/ALDP-9ED167/2020\|EPI_ISL_589441\|20 | .................... | .................... | ..................... | .................... | .................... | .................... |
| M | 7374 | hCoV-19/England/ALDP-9ED4D7/2020\|EPI_ISL_589442\|20 | .................... | .................... | ..................... | .................... | .................... | .................... |
| M | 7375 | hCoV-19/England/ALDP-9ECF9B/2020\|EPI_ISL_589443\|20 | .................... | .................... | ..................... | .................... | .................... | .................... |
| M | 7376 | hCoV-19/England/ALDP-9EE1EE/2020\|EPI_ISL_589444\|20 | .................... | .................... | ..................... | .................... | .................... | .................... |
| M | 7377 | hCoV-19/England/ALDP-9ED079/2020\|EPI_ISL_589445\|20 | .................... | ....A............... | ..................... | .................... | .................... | .................... |
| M | 7378 | hCoV-19/England/ALDP-9ED440/2020\|EPI_ISL_589446\|20 | .................... | .................... | ..................... | .................... | .................... | .................... |
| M | 7379 | hCoV-19/England/ALDP-9EE148/2020\|EPI_ISL_589447\|20 | .................... | .................... | ..................... | .................... | .................... | .................... |
| M | 7380 | hCoV-19/England/ALDP-9E5C31/2020\|EPI_ISL_589448\|20 | .................... | .................... | ..................... | .................... | .................... | .................... |
| M | 7381 | hCoV-19/England/ALDP-9ED5F2/2020\|EPI_ISL_589449\|20 | .................... | .................... | ..................... | .................... | .................... | .................... |
| M | 7382 | hCoV-19/England/ALDP-9ECEDA/2020\|EPI_ISL_589450\|20 | .................... | .................... | ..................... | .................... | .................... | .................... |
| M | 7383 | hCoV-19/England/ALDP-9EE1FD/2020\|EPI_ISL_589451\|20 | .................... | .................... | ..................... | .................... | .................... | .................... |
| M | 7384 | hCoV-19/England/ALDP-9E5949/2020\|EPI_ISL_589452\|20 | .................... | .................... | ..................... | .................... | .................... | .................... |
| M | 7385 | hCoV-19/England/ALDP-9EDB69/2020\|EPI_ISL_589453\|20 | .................... | .................... | ..................... | .................... | .................... | .................... |
| M | 7386 | hCoV-19/England/ALDP-9EE272/2020\|EPI_ISL_589454\|20 | .................... | .................... | ..................... | .................... | .................... | .................... |
| M | 7387 | hCoV-19/England/ALDP-9EDFC7/2020\|EPI_ISL_589455\|20 | .................... | .................... | ..................... | .................... | .................... | .................... |
| M | 7388 | hCoV-19/England/ALDP-9E5BAD/2020\|EPI_ISL_589456\|20 | .................... | .................... | ..................... | .................... | .................... | .................... |
| M | 7389 | hCoV-19/England/ALDP-9ED264/2020\|EPI_ISL_589457\|20 | .................... | .................... | ..................... | .................... | .................... | .................... |
| M | 7390 | hCoV-19/England/ALDP-9E5BDA/2020\|EPI_ISL_589458\|20 | .................... | .................... | ..................... | .................... | .................... | .................... |
| M | 7391 | hCoV-19/England/QEUH-9B5186/2020\|EPI_ISL_590420\|20 | .................... | .A.................. | ..................... | .................... | .................... | ...A................ |
| M | 7392 | hCoV-19/England/QEUH-9B4767/2020\|EPI_ISL_590421\|20 | .................... | .................... | ..................... | .................... | .................... | .................... |
| M | 7393 | hCoV-19/England/ALDP-9B59BB/2020\|EPI_ISL_590422\|20 | .................... | .................... | ..................... | .................... | .................... | .................... |
| M | 7394 | hCoV-19/England/QEUH-9B4785/2020\|EPI_ISL_590424\|20 | .................... | .................... | ..................... | .................... | .................... | .................... |
| M | 7395 | hCoV-19/England/QEUH-9B5265/2020\|EPI_ISL_590425\|20 | .................... | .................... | ..................... | .................... | .................... | .................... |
| M | 7396 | hCoV-19/England/QEUH-9B5308/2020\|EPI_ISL_590426\|20 | .................... | .................... | ..................... | .................... | ....T............... | .................... |
| M | 7397 | hCoV-19/England/QEUH-9B5238/2020\|EPI_ISL_590427\|20 | .................... | .................... | ..................... | .................... | .................... | .................... |
| M | 7398 | hCoV-19/England/QEUH-9B504D/2020\|EPI_ISL_590428\|20 | .................... | .................... | ..................... | .................... | ....T............... | .................... |
| M | 7399 | hCoV-19/England/QEUH-9B4E62/2020\|EPI_ISL_590430\|20 | .................... | .................... | ..................... | .................... | .................... | .................... |
| M | 7400 | hCoV-19/England/QEUH-9B47D0/2020\|EPI_ISL_590431\|20 | .................... | .................... | ..................... | .................... | .................... | .................... |
| M | 7401 | hCoV-19/Wales/QEUH-9B49BC/2020\|EPI_ISL_590432\|2020 | .................... | .................... | ..................... | .................... | .................... | .................... |
| M | 7402 | hCoV-19/England/QEUH-9B50A7/2020\|EPI_ISL_590434\|20 | .................... | .................... | ..................... | .................... | .................... | .................... |
| M | 7403 | hCoV-19/England/QEUH-9B52A1/2020\|EPI_ISL_590435\|20 | .................... | .................... | ..................... | .................... | .................... | .................... |
| M | 7404 | hCoV-19/England/QEUH-9B5405/2020\|EPI_ISL_590436\|20 | .................... | .................... | ..................... | .................... | .................... | .................... |
| M | 7405 | hCoV-19/England/QEUH-9B56A5/2020\|EPI_ISL_590437\|20 | .................... | .................... | ..................... | .................... | .................... | .................... |
| M | 7406 | hCoV-19/England/ALDP-9B5A4F/2020\|EPI_ISL_590438\|20 | .................... | .................... | ..................... | .................... | .................... | .................... |
| M | 7407 | hCoV-19/England/QEUH-9B563C/2020\|EPI_ISL_590439\|20 | .................... | .................... | ..................... | .................... | .................... | .................... |
| M | 7408 | hCoV-19/England/QEUH-9B798C/2020\|EPI_ISL_590482\|20 | .................... | .................... | ..................... | .................... | ....T............... | .................... |
| M | 7409 | hCoV-19/England/QEUH-9B7C56/2020\|EPI_ISL_590483\|20 | .................... | .A.................. | ..................... | .................... | .................... | .................... |
| M | 7410 | hCoV-19/England/QEUH-9B7931/2020\|EPI_ISL_590484\|20 | .................... | .................... | ..................... | .................... | .................... | .................... |
| M | 7411 | hCoV-19/England/QEUH-9B789E/2020\|EPI_ISL_590485\|20 | .................... | .................... | ..................... | .................... | .................... | .................... |
| M | 7412 | hCoV-19/England/QEUH-9B7C47/2020\|EPI_ISL_590486\|20 | .................... | .A.................. | ..................... | .................... | .................... | .................... |
| M | 7413 | hCoV-19/Wales/QEUH-9B7D17/2020\|EPI_ISL_590487\|2020 | .................... | .A.................. | ..................... | .................... | .................... | .................... |
| M | 7414 | hCoV-19/England/QEUH-9B7421/2020\|EPI_ISL_590488\|20 | .................... | .................... | ..................... | .................... | .................... | .................... |
| M | 7415 | hCoV-19/NorthernIreland/QEUH-9B7412/2020\|EPI_ISL_5 | .................... | .................... | ..................... | .................... | .................... | .................... |
| M | 7416 | hCoV-19/England/QEUH-9B7BFF/2020\|EPI_ISL_590490\|20 | .................... | .................... | ..................... | .................... | .................... | .................... |
| M | 7417 | hCoV-19/Wales/QEUH-9B70D2/2020\|EPI_ISL_590491\|2020 | .................... | .................... | ..................... | .................... | .................... | .................... |
| M | 7418 | hCoV-19/England/QEUH-9B7430/2020\|EPI_ISL_590492\|20 | .................... | .................... | ..................... | .................... | .................... | .................... |
| M | 7419 | hCoV-19/England/QEUH-9B8077/2020\|EPI_ISL_590494\|20 | .................... | .................... | ..................... | .................... | .................... | .................... |
| M | 7420 | hCoV-19/England/QEUH-9B7676/2020\|EPI_ISL_590495\|20 | .................... | .................... | ..................... | .................... | .................... | .................... |
| M | 7421 | hCoV-19/England/QEUH-9B712A/2020\|EPI_ISL_590496\|20 | .................... | .................... | ..................... | .................... | .................... | ...............A.... |
| M | 7422 | hCoV-19/England/QEUH-9B7773/2020\|EPI_ISL_590498\|20 | .................... | .................... | ..................... | .................... | .................... | .................... |
| M | 7423 | hCoV-19/England/QEUH-9B79F5/2020\|EPI_ISL_590499\|20 | .................... | .................... | ..................... | .................... | .................... | .................... |
| M | 7424 | hCoV-19/France/OCC-32/2020\|EPI_ISL_591099\|2020-06- | .................... | .................... | ..................... | .................... | .................... | .................... |
| M | 7425 | hCoV-19/England/QEUH-9FAE51/2020\|EPI_ISL_588100\|20 | .................... | .................... | ..................... | .................... | .................... | .................... |
| M | 7426 | hCoV-19/England/QEUH-9FA53E/2020\|EPI_ISL_588101\|20 | .................... | .................... | ..................... | .................... | .................... | .................... |
| M | 7427 | hCoV-19/England/QEUH-9FB087/2020\|EPI_ISL_588102\|20 | .................... | .................... | ..................... | .................... | .................... | .................... |
| M | 7428 | hCoV-19/England/QEUH-9FA8AE/2020\|EPI_ISL_588103\|20 | .................... | .................... | ..................... | .................... | .................... | .................... |
| M | 7429 | hCoV-19/England/QEUH-9FB1B1/2020\|EPI_ISL_588104\|20 | .................... | .................... | ..................... | .................... | .................... | .................... |
| M | 7430 | hCoV-19/England/QEUH-9FA729/2020\|EPI_ISL_588105\|20 | .................... | .................... | ..................... | .................... | .................... | .................... |
| M | 7431 | hCoV-19/England/QEUH-9FB272/2020\|EPI_ISL_588129\|20 | .................... | .................... | ..................... | .................... | ....T............... | .................... |
| M | 7432 | hCoV-19/England/QEUH-9FA3F8/2020\|EPI_ISL_588130\|20 | .................... | .................... | ..................... | .................... | .................... | .................... |
| M | 7433 | hCoV-19/England/QEUH-9FB0F0/2020\|EPI_ISL_588132\|20 | .................... | .................... | ..................... | .................... | .................... | .................... |
| M | 7434 | hCoV-19/England/QEUH-9FB166/2020\|EPI_ISL_588133\|20 | .................... | .................... | ..................... | .................... | .................... | .................... |
| M | 7435 | hCoV-19/England/QEUH-9FA8CC/2020\|EPI_ISL_588134\|20 | .................... | .................... | ..................... | .................... | ......T............. | .................... |
| M | 7436 | hCoV-19/England/QEUH-9FACEE/2020\|EPI_ISL_588135\|20 | .................... | .................... | ..................... | .................... | .................... | .................... |
| M | 7437 | hCoV-19/England/QEUH-9FB157/2020\|EPI_ISL_588136\|20 | .................... | .................... | ..................... | .................... | .................... | .................... |
| M | 7438 | hCoV-19/England/QEUH-9FAA5D/2020\|EPI_ISL_588138\|20 | .................... | .................... | ..................... | .................... | .................... | .................... |
| M | 7439 | hCoV-19/England/QEUH-9FAECA/2020\|EPI_ISL_588139\|20 | .................... | .................... | ..................... | .................... | .................... | .................... |
| M | 7440 | hCoV-19/England/QEUH-9FAB87/2020\|EPI_ISL_588140\|20 | .................... | .................... | ..................... | .................... | .................... | .................... |
| M | 7441 | hCoV-19/England/QEUH-9FB324/2020\|EPI_ISL_588141\|20 | .................... | .................... | ..................... | .................... | ....T............... | .................... |
| M | 7442 | hCoV-19/England/QEUH-9FB2EB/2020\|EPI_ISL_588142\|20 | .................... | .................... | ..................... | .................... | .................... | .................... |
| M | 7443 | hCoV-19/England/QEUH-9FA7FC/2020\|EPI_ISL_588143\|20 | .................... | .A.................. | ..................... | .................... | .................... | ...A................ |
| M | 7444 | hCoV-19/England/QEUH-9FA440/2020\|EPI_ISL_588145\|20 | .................... | .................... | ..................... | .................... | .................... | .................... |
| M | 7445 | hCoV-19/England/QEUH-9FB139/2020\|EPI_ISL_588146\|20 | .................... | .................... | ..................... | .................... | .................... | .................... |
| M | 7446 | hCoV-19/England/QEUH-9FA8F9/2020\|EPI_ISL_588147\|20 | .................... | .................... | ..................... | .................... | .................... | .................... |
| M | 7447 | hCoV-19/England/QEUH-9FABA5/2020\|EPI_ISL_588148\|20 | .................... | .................... | ..................... | .................... | .................... | .................... |
| M | 7448 | hCoV-19/England/QEUH-9FA64A/2020\|EPI_ISL_588149\|20 | .................... | .................... | ..................... | .................... | .................... | .................... |
| M | 7449 | hCoV-19/England/QEUH-9FA422/2020\|EPI_ISL_588150\|20 | .................... | .................... | ..................... | .................... | .................... | .................... |
| M | 7450 | hCoV-19/England/QEUH-9FAAC6/2020\|EPI_ISL_588151\|20 | .................... | .................... | ..................... | .................... | .................... | .................... |
| M | 7451 | hCoV-19/England/QEUH-9FAE15/2020\|EPI_ISL_588152\|20 | .................... | .................... | ..................... | .................... | .................... | .................... |

| | | | | | | | | |
|---|---|---|---|---|---|---|---|---|
| M | 7452 | hCoV-19/England/QEUH-9FB02D/2020\|EPI_ISL_588153\|20 | ···················· | ···················· | ····················· | ···················· | ···················· | ···················· |
| M | 7453 | hCoV-19/England/QEUH-9FABF0/2020\|EPI_ISL_588154\|20 | ···················· | ···················· | ····················· | ···················· | ···················· | ···················· |
| M | 7454 | hCoV-19/England/QEUH-9FAE8E/2020\|EPI_ISL_588155\|20 | ···················· | ···················· | ····················· | ···················· | ···················· | ···················· |
| M | 7455 | hCoV-19/England/QEUH-9FA5E3/2020\|EPI_ISL_588156\|20 | ···················· | ···················· | ····················· | ···················· | ···················· | ···················· |
| M | 7456 | hCoV-19/England/QEUH-9FB4E5/2020\|EPI_ISL_588157\|20 | ···················· | ···················· | ····················· | ···················· | ···················· | ···················· |
| M | 7457 | hCoV-19/England/QEUH-9FADDC/2020\|EPI_ISL_588158\|20 | ···················· | ···················· | ····················· | ···················· | ···················· | ···················· |
| M | 7458 | hCoV-19/England/QEUH-9FACB1/2020\|EPI_ISL_588159\|20 | ···················· | ···················· | ····················· | ···················· | ···················· | ···················· |
| M | 7459 | hCoV-19/England/QEUH-9FB360/2020\|EPI_ISL_588160\|20 | ···················· | ···················· | ····················· | ···················· | ···················· | ···················· |
| M | 7460 | hCoV-19/England/QEUH-9FA7B0/2020\|EPI_ISL_588161\|20 | ···················· | ·A·················· | ····················· | ···················· | ···················· | ···················· |
| M | 7461 | hCoV-19/England/QEUH-9FB0D2/2020\|EPI_ISL_588162\|20 | ···················· | ···················· | ····················· | ···················· | ···················· | ···················· |
| M | 7462 | hCoV-19/England/QEUH-9FAFD6/2020\|EPI_ISL_588163\|20 | ···················· | ···················· | ····················· | ···················· | ···················· | ···················· |
| M | 7463 | hCoV-19/England/QEUH-9FAD18/2020\|EPI_ISL_588164\|20 | ···················· | ···G················ | ····················· | ···················· | ···················· | ···················· |
| M | 7464 | hCoV-19/England/QEUH-9FB0C3/2020\|EPI_ISL_588165\|20 | ···················· | ···················· | ····················· | ···················· | ···················· | ···················· |
| M | 7465 | hCoV-19/England/QEUH-9FB56A/2020\|EPI_ISL_588166\|20 | ···················· | ···················· | ····················· | ···················· | ···················· | ···················· |
| M | 7466 | hCoV-19/England/QEUH-9FAF12/2020\|EPI_ISL_588167\|20 | ···················· | ···················· | ····················· | ···················· | ···················· | ···················· |
| M | 7467 | hCoV-19/England/QEUH-9FAAA8/2020\|EPI_ISL_588168\|20 | ···················· | ·A·················· | ····················· | ···················· | ···················· | ···················· |
| M | 7468 | hCoV-19/England/QEUH-9FA510/2020\|EPI_ISL_588169\|20 | ···················· | ·A·················· | ····················· | ···················· | ···················· | ···················· |
| M | 7469 | hCoV-19/England/QEUH-9FAC0C/2020\|EPI_ISL_588200\|20 | ···················· | ···················· | ····················· | ···················· | ···················· | ···················· |
| M | 7470 | hCoV-19/England/QEUH-9FA57A/2020\|EPI_ISL_588201\|20 | ···················· | ···················· | ····················· | ···················· | ···················· | ···················· |
| M | 7471 | hCoV-19/England/QEUH-9FA352/2020\|EPI_ISL_588202\|20 | ···················· | ···················· | ····················· | ···················· | ···················· | ···················· |
| M | 7472 | hCoV-19/England/QEUH-9FB290/2020\|EPI_ISL_588203\|20 | ···················· | ···················· | ····················· | ···················· | ···················· | ···················· |
| M | 7473 | hCoV-19/England/QEUH-9FAC1B/2020\|EPI_ISL_588204\|20 | ···················· | ···················· | ····················· | ···················· | ···················· | ···················· |
| M | 7474 | hCoV-19/England/ALDP-9ED6C2/2020\|EPI_ISL_589422\|20 | ···················· | ···················· | ····················· | ···················· | ···················· | ···················· |
| M | 7475 | hCoV-19/England/ALDP-9EE0A5/2020\|EPI_ISL_589424\|20 | ···················· | ···················· | ····················· | ···················· | ···················· | ···················· |
| M | 7476 | hCoV-19/England/ALDP-9ECE52/2020\|EPI_ISL_589425\|20 | ···················· | ···················· | ····················· | ···················· | ···················· | ···················· |
| M | 7477 | hCoV-19/England/ALDP-9E5C22/2020\|EPI_ISL_589426\|20 | ···················· | ···················· | ····················· | ···················· | ···················· | ···················· |
| M | 7478 | hCoV-19/England/ALDP-9EDED9/2020\|EPI_ISL_589427\|20 | ···················· | ···················· | ····················· | ···················· | ···················· | ···················· |
| M | 7479 | hCoV-19/England/ALDP-9ED307/2020\|EPI_ISL_589428\|20 | ···················· | ···················· | ····················· | ···················· | ···················· | ···················· |
| M | 7480 | hCoV-19/England/ALDP-9E57D6/2020\|EPI_ISL_589429\|20 | ···················· | ···················· | ····················· | ···················· | ···················· | ···················· |
| M | 7481 | hCoV-19/England/MILK-9ECC2B/2020\|EPI_ISL_589430\|20 | ···················· | ···················· | ····················· | ···················· | ···················· | ···················· |
| M | 7482 | hCoV-19/England/ALDP-9E5976/2020\|EPI_ISL_589431\|20 | ···················· | ···················· | ····················· | ···················· | ···················· | ···················· |
| M | 7483 | hCoV-19/England/ALDP-9E5C7D/2020\|EPI_ISL_589432\|20 | ···················· | ···················· | ····················· | ···················· | ···················· | ···················· |
| M | 7484 | hCoV-19/England/ALDP-9EE333/2020\|EPI_ISL_589433\|20 | ···················· | ···················· | ····················· | ···················· | ···················· | ···················· |
| M | 7485 | hCoV-19/England/ALDP-9E5A37/2020\|EPI_ISL_589434\|20 | ···················· | ···················· | ····················· | ···················· | ···················· | ···················· |
| M | 7486 | hCoV-19/England/ALDP-9ECEBC/2020\|EPI_ISL_589435\|20 | ···················· | ···················· | ····················· | ···················· | ···················· | ···················· |
| M | 7487 | hCoV-19/England/ALDP-9E5BCB/2020\|EPI_ISL_589436\|20 | ···················· | ···················· | ····················· | ···················· | ···················· | ···················· |
| M | 7488 | hCoV-19/England/ALDP-9E5A0A/2020\|EPI_ISL_589459\|20 | ···················· | ···················· | ····················· | ···················· | ···················· | ···················· |
| M | 7489 | hCoV-19/England/ALDP-9ECD82/2020\|EPI_ISL_589460\|20 | ···················· | ···················· | ····················· | ···················· | ···················· | ···················· |
| M | 7490 | hCoV-19/England/ALDP-9EDBA5/2020\|EPI_ISL_589462\|20 | ···················· | ···················· | ····················· | ···················· | ···················· | ···················· |
| M | 7491 | hCoV-19/England/ALDP-9EE12A/2020\|EPI_ISL_589463\|20 | ···················· | ···················· | ····················· | ···················· | ···················· | ···················· |
| M | 7492 | hCoV-19/England/ALDP-9ED1FE/2020\|EPI_ISL_589464\|20 | ···················· | ···················· | ····················· | ···················· | ···················· | ···················· |
| M | 7493 | hCoV-19/England/ALDP-9ED2CE/2020\|EPI_ISL_589465\|20 | ···················· | ···················· | ····················· | ···················· | ···················· | ···················· |
| M | 7494 | hCoV-19/England/ALDP-9E592B/2020\|EPI_ISL_589466\|20 | ···················· | ···················· | ····················· | ···················· | ···················· | ···················· |
| M | 7495 | hCoV-19/England/ALDP-9ED45F/2020\|EPI_ISL_589468\|20 | ···················· | ···················· | ····················· | ···················· | ···················· | ···················· |
| M | 7496 | hCoV-19/England/ALDP-9EE0C3/2020\|EPI_ISL_589469\|20 | ···················· | ···················· | ····················· | ···················· | ···················· | ···················· |
| M | 7497 | hCoV-19/England/ALDP-9ED02E/2020\|EPI_ISL_589470\|20 | ···················· | ···················· | ····················· | ···················· | ···················· | ···················· |
| M | 7498 | hCoV-19/England/ALDP-9ED10D/2020\|EPI_ISL_589471\|20 | ···················· | ···················· | ····················· | ···················· | ···················· | ···················· |
| M | 7499 | hCoV-19/England/ALDP-9E56F7/2020\|EPI_ISL_589472\|20 | ···················· | ···················· | ····················· | ···················· | ···················· | ···················· |
| M | 7500 | hCoV-19/England/ALDP-9E5A19/2020\|EPI_ISL_589473\|20 | ···················· | ···················· | ····················· | ···················· | ···················· | ···················· |
| M | 7501 | hCoV-19/England/ALDP-9E5A55/2020\|EPI_ISL_589474\|20 | ···················· | ···················· | ····················· | ···················· | ···················· | ···················· |
| M | 7502 | hCoV-19/England/ALDP-9EDF5E/2020\|EPI_ISL_589476\|20 | ···················· | ···················· | ····················· | ···················· | ···················· | ···················· |
| M | 7503 | hCoV-19/England/ALDP-9E593A/2020\|EPI_ISL_589478\|20 | ···················· | ···················· | ····················· | ···················· | ···················· | ···················· |
| M | 7504 | hCoV-19/England/ALDP-9E5BBC/2020\|EPI_ISL_589479\|20 | ···················· | ···················· | ····················· | ···················· | ···················· | ···················· |
| M | 7505 | hCoV-19/England/ALDP-9EDDCD/2020\|EPI_ISL_589480\|20 | ···················· | ···················· | ····················· | ···················· | ···················· | ···················· |
| M | 7506 | hCoV-19/England/ALDP-9E591C/2020\|EPI_ISL_589481\|20 | ···················· | ···················· | ····················· | ···················· | ···················· | ···················· |
| M | 7507 | hCoV-19/England/ALDP-9E59EF/2020\|EPI_ISL_589482\|20 | ···················· | ···················· | ····················· | ···················· | ···················· | ···················· |
| M | 7508 | hCoV-19/England/ALDP-9ECE8F/2020\|EPI_ISL_589483\|20 | ···················· | ···················· | ····················· | ···················· | ···················· | ···················· |
| M | 7509 | hCoV-19/England/ALDP-9E585B/2020\|EPI_ISL_589484\|20 | ···················· | ···················· | ····················· | ···················· | ···················· | ···················· |
| M | 7510 | hCoV-19/England/ALDP-9ED64A/2020\|EPI_ISL_589485\|20 | ···················· | ···················· | ····················· | ···················· | ···················· | ···················· |
| M | 7511 | hCoV-19/England/ALDP-9EDDEB/2020\|EPI_ISL_589486\|20 | ···················· | ···················· | ····················· | ···················· | ···················· | ···················· |
| M | 7512 | hCoV-19/England/ALDP-9ED325/2020\|EPI_ISL_589487\|20 | ···················· | ···················· | ····················· | ···················· | ···················· | ···················· |
| M | 7513 | hCoV-19/England/ALDP-9E583D/2020\|EPI_ISL_589488\|20 | ···················· | ···················· | ····················· | ···················· | ···················· | ···················· |
| M | 7514 | hCoV-19/England/ALDP-9EE2FA/2020\|EPI_ISL_589489\|20 | ···················· | ···················· | ····················· | ···················· | ···················· | ···················· |
| M | 7515 | hCoV-19/England/ALDP-9ED282/2020\|EPI_ISL_589490\|20 | ···················· | ···················· | ····················· | ···················· | ···················· | ···················· |
| M | 7516 | hCoV-19/England/ALDP-9E5D10/2020\|EPI_ISL_589491\|20 | ···················· | ···················· | ····················· | ···················· | ···················· | ···················· |
| M | 7517 | hCoV-19/England/ALDP-9EE175/2020\|EPI_ISL_589492\|20 | ···················· | ···················· | ····················· | ···················· | ···················· | ···················· |
| M | 7518 | hCoV-19/England/MILK-9ECCC1/2020\|EPI_ISL_589493\|20 | ···················· | ···················· | ····················· | ···················· | ···················· | ···················· |
| M | 7519 | hCoV-19/England/ALDP-9E56D9/2020\|EPI_ISL_589494\|20 | ···················· | ···················· | ····················· | ···················· | ···················· | ···················· |
| M | 7520 | hCoV-19/England/ALDP-9EDB78/2020\|EPI_ISL_589495\|20 | ···················· | ···················· | ····················· | ···················· | ···················· | ···················· |
| M | 7521 | hCoV-19/England/ALDP-9EE01E/2020\|EPI_ISL_589496\|20 | ···················· | ···················· | ····················· | ···················· | ···················· | ···················· |
| M | 7522 | hCoV-19/England/ALDP-9E5BE9/2020\|EPI_ISL_589497\|20 | ···················· | ···················· | ····················· | ···················· | ···················· | ···················· |
| M | 7523 | hCoV-19/England/ALDP-9ED5B6/2020\|EPI_ISL_589498\|20 | ···················· | ···················· | ····················· | ···················· | ···················· | ···················· |
| M | 7524 | hCoV-19/England/ALDP-9E5B70/2020\|EPI_ISL_589499\|20 | ···················· | ···················· | ····················· | ···················· | ···················· | ···················· |
| M | 7525 | hCoV-19/England/ALDP-9ED1B2/2020\|EPI_ISL_589514\|20 | ···················· | ···················· | ····················· | ···················· | ···················· | ···················· |
| M | 7526 | hCoV-19/England/ALDP-9ED1C1/2020\|EPI_ISL_589515\|20 | ···················· | ···················· | ····················· | ···················· | ···················· | ···················· |
| M | 7527 | hCoV-19/England/ALDP-9ED844/2020\|EPI_ISL_589516\|20 | ···················· | ···················· | ····················· | ···················· | ···················· | ···················· |
| M | 7528 | hCoV-19/England/ALDP-9ECD19/2020\|EPI_ISL_589517\|20 | ···················· | ···················· | ····················· | ···················· | ···················· | ···················· |
| M | 7529 | hCoV-19/England/ALDP-9EE087/2020\|EPI_ISL_589518\|20 | ···················· | ···················· | ····················· | ···················· | ···················· | ···················· |
| M | 7530 | hCoV-19/England/MILK-9ECBF1/2020\|EPI_ISL_589519\|20 | ···················· | ···················· | ····················· | ···················· | ···················· | ···················· |
| M | 7531 | hCoV-19/England/MILK-9ECD73/2020\|EPI_ISL_589520\|20 | ···················· | ···················· | ····················· | ···················· | ···················· | ···················· |

| | | | | | | | |
|---|---|---|---|---|---|---|---|
| M | 7532 hCoV-19/England/MILK-9ECBD3/2020\|EPI_ISL_589521\|20 | .................... | .................... | .................... | .................... | .................... | .................... |
| M | 7533 hCoV-19/England/ALDP-9E59C1/2020\|EPI_ISL_589522\|20 | .................... | .................... | .................... | .................... | .................... | .................... |
| M | 7534 hCoV-19/England/ALDP-9E5A82/2020\|EPI_ISL_589523\|20 | .................... | .................... | .................... | .................... | .................... | .................... |
| M | 7535 hCoV-19/England/ALDP-9EDEF7/2020\|EPI_ISL_589524\|20 | .................... | .................... | .................... | .................... | .................... | .................... |
| M | 7536 hCoV-19/England/ALDP-9EE0F0/2020\|EPI_ISL_589525\|20 | .................... | .................... | .................... | .................... | .................... | .................... |
| M | 7537 hCoV-19/England/ALDP-9ED0D3/2020\|EPI_ISL_589527\|20 | .................... | .................... | .................... | .................... | .................... | .................... |
| M | 7538 hCoV-19/England/ALDP-9ECCFE/2020\|EPI_ISL_589528\|20 | .................... | .................... | .................... | .................... | .................... | .................... |
| M | 7539 hCoV-19/England/ALDP-9ECE70/2020\|EPI_ISL_589529\|20 | .................... | .................... | .................... | .................... | .................... | .................... |
| M | 7540 hCoV-19/England/ALDP-9E5712/2020\|EPI_ISL_589530\|20 | .................... | .................... | .................... | .................... | .................... | .................... |
| M | 7541 hCoV-19/England/ALDP-9E5B8F/2020\|EPI_ISL_589531\|20 | .................... | .................... | .................... | .................... | .................... | .................... |
| M | 7542 hCoV-19/England/ALDP-9E57B8/2020\|EPI_ISL_589532\|20 | .................... | .R.................. | .................... | .................... | .................... | .................... |
| M | 7543 hCoV-19/England/ALDP-9E57E5/2020\|EPI_ISL_589533\|20 | .................... | .................... | .................... | .................... | .................... | .................... |
| M | 7544 hCoV-19/England/ALDP-9ED5D4/2020\|EPI_ISL_589534\|20 | .................... | .................... | .................... | .................... | .................... | .................... |
| M | 7545 hCoV-19/England/ALDP-9EE2EB/2020\|EPI_ISL_589535\|20 | .................... | .................... | .................... | .................... | .................... | .................... |
| M | 7546 hCoV-19/England/QEUH-9B46B5/2020\|EPI_ISL_590400\|20 | .................... | .................... | .................... | .................... | .................... | .................... |
| M | 7547 hCoV-19/England/QEUH-9B558A/2020\|EPI_ISL_590401\|20 | .................... | .................... | .................... | .................... | .................... | .................... |
| M | 7548 hCoV-19/England/ALDP-9B5AA9/2020\|EPI_ISL_590403\|20 | .................... | .................... | .................... | .................... | .................... | .................... |
| M | 7549 hCoV-19/England/QEUH-9B456D/2020\|EPI_ISL_590404\|20 | .................... | .................... | .................... | .................... | .................... | .................... |
| M | 7550 hCoV-19/England/ALDP-9B5845/2020\|EPI_ISL_590405\|20 | .................... | .................... | .................... | .................... | .................... | .................... |
| M | 7551 hCoV-19/England/QEUH-9B5669/2020\|EPI_ISL_590406\|20 | .................... | .................... | .................... | .................... | .................... | .................... |
| M | 7552 hCoV-19/England/QEUH-9B53EA/2020\|EPI_ISL_590407\|20 | .................... | .................... | .................... | .................... | .................... | .................... |
| M | 7553 hCoV-19/England/QEUH-9B4D83/2020\|EPI_ISL_590408\|20 | .................... | .................... | .................... | .................... | .................... | .................... |
| M | 7554 hCoV-19/England/QEUH-9B4DB0/2020\|EPI_ISL_590409\|20 | .................... | .................... | .................... | .................... | .................... | .................... |
| M | 7555 hCoV-19/England/QEUH-9B5502/2020\|EPI_ISL_590410\|20 | .................... | .................... | .................... | .................... | .................... | .................... |
| M | 7556 hCoV-19/England/QEUH-9B561E/2020\|EPI_ISL_590411\|20 | .................... | .................... | .................... | .................... | .................... | .................... |
| M | 7557 hCoV-19/England/QEUH-9B5326/2020\|EPI_ISL_590412\|20 | .................... | .................... | .................... | .................... | .................... | .................... |
| M | 7558 hCoV-19/England/QEUH-9B4907/2020\|EPI_ISL_590413\|20 | .................... | .................... | .................... | .................... | .................... | .................... |
| M | 7559 hCoV-19/England/QEUH-9B48A0/2020\|EPI_ISL_590414\|20 | .................... | .................... | .................... | .................... | .................... | .................... |
| M | 7560 hCoV-19/England/QEUH-9B4E35/2020\|EPI_ISL_590415\|20 | .................... | .................... | .................... | .................... | .................... | .................... |
| M | 7561 hCoV-19/England/QEUH-9B4C2C/2020\|EPI_ISL_590416\|20 | .................... | .................... | .................... | .................... | .................... | .................... |
| M | 7562 hCoV-19/England/QEUH-9B49F8/2020\|EPI_ISL_590417\|20 | .................... | .................... | .................... | .................... | .................... | .................... |
| M | 7563 hCoV-19/England/QEUH-9B5344/2020\|EPI_ISL_590418\|20 | .................... | .................... | .................... | .................... | .................... | .................... |
| M | 7564 hCoV-19/England/QEUH-9B5247/2020\|EPI_ISL_590440\|20 | .................... | .................... | .................... | .................... | .................... | .................... |
| M | 7565 hCoV-19/NorthernIreland/QEUH-9B46D3/2020\|EPI_ISL_5 | .................... | .................... | .................... | .................... | .................... | .................... |
| M | 7566 hCoV-19/England/QEUH-9B4C1D/2020\|EPI_ISL_590444\|20 | .................... | .................... | .................... | .................... | .................... | .................... |
| M | 7567 hCoV-19/England/QEUH-9B4E08/2020\|EPI_ISL_590445\|20 | .................... | .................... | .................... | .................... | .................... | .................... |
| M | 7568 hCoV-19/England/ALDP-9B59CA/2020\|EPI_ISL_590446\|20 | .................... | .A.................. | .................... | .................... | .................... | .................... |
| M | 7569 hCoV-19/England/QEUH-9B4B3E/2020\|EPI_ISL_590447\|20 | .................... | .................... | .................... | .................... | .................... | .................... |
| M | 7570 hCoV-19/England/QEUH-9B557B/2020\|EPI_ISL_590449\|20 | .................... | .A.................. | .................... | .................... | .................... | .................... |
| M | 7571 hCoV-19/England/QEUH-9B4A40/2020\|EPI_ISL_590450\|20 | .................... | .................... | .................... | .................... | .................... | .................... |
| M | 7572 hCoV-19/England/QEUH-9B5511/2020\|EPI_ISL_590451\|20 | .................... | .................... | .................... | .................... | .................... | .................... |
| M | 7573 hCoV-19/England/QEUH-9B49AD/2020\|EPI_ISL_590454\|20 | .................... | .................... | .................... | .................... | .................... | .................... |
| M | 7574 hCoV-19/England/QEUH-9B4D47/2020\|EPI_ISL_590455\|20 | .................... | .................... | .................... | .................... | .................... | .................... |
| M | 7575 hCoV-19/England/QEUH-9B562D/2020\|EPI_ISL_590456\|20 | .................... | .................... | .................... | .................... | .................... | .................... |
| M | 7576 hCoV-19/NorthernIreland/QEUH-9B459A/2020\|EPI_ISL_5 | .................... | .................... | .................... | .................... | .................... | .................... |
| M | 7577 hCoV-19/England/QEUH-9B4AAA/2020\|EPI_ISL_590458\|20 | .................... | .................... | .................... | .................... | .................... | .................... |
| M | 7578 hCoV-19/England/QEUH-9B5423/2020\|EPI_ISL_590459\|20 | .................... | .................... | .................... | .................... | .................... | .................... |
| M | 7579 hCoV-19/England/QEUH-9B51D1/2020\|EPI_ISL_590460\|20 | .................... | .................... | .................... | .................... | .................... | .................... |
| M | 7580 hCoV-19/England/QEUH-9B5432/2020\|EPI_ISL_590461\|20 | .................... | .................... | .................... | .................... | .................... | .................... |
| M | 7581 hCoV-19/England/ALDP-9B5A8B/2020\|EPI_ISL_590462\|20 | .................... | .................... | .................... | .................... | .................... | .................... |
| M | 7582 hCoV-19/England/QEUH-9B77DD/2020\|EPI_ISL_590463\|20 | .................... | .................... | .................... | .................... | ....T............... | .................... |
| M | 7583 hCoV-19/England/QEUH-9B7685/2020\|EPI_ISL_590464\|20 | .................... | .................... | .................... | .................... | .................... | .................... |
| M | 7584 hCoV-19/England/QEUH-9B7A2F/2020\|EPI_ISL_590465\|20 | .................... | .................... | .................... | .................... | .................... | .................... |
| M | 7585 hCoV-19/England/QEUH-9B81A1/2020\|EPI_ISL_590466\|20 | .................... | .................... | .................... | .................... | .................... | .................... |
| M | 7586 hCoV-19/England/QEUH-9B7ED8/2020\|EPI_ISL_590467\|20 | .................... | .................... | .................... | .................... | .................... | .................... |
| M | 7587 hCoV-19/England/QEUH-9B6EAC/2020\|EPI_ISL_590468\|20 | .................... | .................... | .................... | .................... | .................... | .................... |
| M | 7588 hCoV-19/England/QEUH-9B72AF/2020\|EPI_ISL_590469\|20 | .................... | .................... | .................... | .................... | .................... | .................... |
| M | 7589 hCoV-19/England/QEUH-9B7B59/2020\|EPI_ISL_590470\|20 | .................... | .................... | .................... | .................... | .................... | .................... |
| M | 7590 hCoV-19/England/QEUH-9B7D9F/2020\|EPI_ISL_590471\|20 | .................... | .................... | .................... | .................... | .................... | .................... |
| M | 7591 hCoV-19/England/QEUH-9B7C1A/2020\|EPI_ISL_590472\|20 | .................... | .................... | .................... | .................... | .................... | .................... |
| M | 7592 hCoV-19/England/QEUH-9B7D08/2020\|EPI_ISL_590473\|20 | .................... | .................... | .................... | .................... | .................... | .................... |
| M | 7593 hCoV-19/England/QEUH-9B7B77/2020\|EPI_ISL_590474\|20 | .................... | .................... | .................... | .................... | .................... | .................... |
| M | 7594 hCoV-19/England/QEUH-9B7069/2020\|EPI_ISL_590475\|20 | .................... | .................... | .................... | .................... | .................... | .................... |
| M | 7595 hCoV-19/England/QEUH-9B7AA7/2020\|EPI_ISL_590476\|20 | .................... | .................... | .................... | .................... | .................... | .................... |
| M | 7596 hCoV-19/England/QEUH-9B7940/2020\|EPI_ISL_590477\|20 | .................... | .................... | .................... | .................... | .................... | .................... |
| M | 7597 hCoV-19/England/QEUH-9B701E/2020\|EPI_ISL_590478\|20 | .................... | .................... | .................... | .................... | .................... | .................... |
| M | 7598 hCoV-19/England/QEUH-9B7FB7/2020\|EPI_ISL_590479\|20 | .................... | .................... | .................... | .................... | .................... | .................... |
| M | 7599 hCoV-19/England/QEUH-9B7EF6/2020\|EPI_ISL_590480\|20 | .................... | .................... | .................... | .................... | ....T............... | .................... |
| M | 7600 hCoV-19/England/QEUH-9B7342/2020\|EPI_ISL_590500\|20 | .................... | .................... | .................... | .................... | .................... | .................... |
| M | 7601 hCoV-19/England/QEUH-9B75E2/2020\|EPI_ISL_590501\|20 | .................... | .................... | .................... | .................... | .................... | .................... |
| M | 7602 hCoV-19/England/QEUH-9B7218/2020\|EPI_ISL_590502\|20 | .................... | .................... | .................... | .................... | .................... | .................... |
| M | 7603 hCoV-19/England/QEUH-9B7CFC/2020\|EPI_ISL_590503\|20 | .................... | .................... | .................... | .................... | .................... | .................... |
| M | 7604 hCoV-19/England/QEUH-9B74E5/2020\|EPI_ISL_590504\|20 | .................... | .A.................. | .................... | .................... | .................... | ...A................ |
| M | 7605 hCoV-19/England/QEUH-9B79B9/2020\|EPI_ISL_590505\|20 | .................... | .................... | .................... | .................... | .................... | .................... |
| M | 7606 hCoV-19/England/QEUH-9B7A5C/2020\|EPI_ISL_590506\|20 | .................... | .................... | .................... | .................... | .................... | .................... |
| M | 7607 hCoV-19/England/QEUH-9B7579/2020\|EPI_ISL_590508\|20 | .................... | .................... | .................... | .................... | .................... | .................... |
| M | 7608 hCoV-19/England/QEUH-9B75F1/2020\|EPI_ISL_590509\|20 | .................... | .................... | .................... | .................... | .................... | .................... |
| M | 7609 hCoV-19/England/QEUH-9B78DA/2020\|EPI_ISL_590510\|20 | .................... | .................... | .................... | .................... | .................... | .................... |
| M | 7610 hCoV-19/England/QEUH-9B7B4A/2020\|EPI_ISL_590511\|20 | .................... | .................... | .................... | .................... | ......T............. | .................... |
| M | 7611 hCoV-19/England/QEUH-9B74F4/2020\|EPI_ISL_590516\|20 | .................... | .................... | .................... | .................... | .................... | .................... |

| | | | | | | | | | |
|---|---|---|---|---|---|---|---|---|---|
| M | 7612 | hCoV-19/England/QEUH-9B801D/2020\|EPI_ISL_590517\|20 | ···················· | ···················· | ····················· | ···················· | ···················· | ···················· | |
| M | 7613 | hCoV-19/England/QEUH-9B7EE7/2020\|EPI_ISL_590518\|20 | ···················· | ···················· | ····················· | ···················· | ···················· | ···················· | |
| M | 7614 | hCoV-19/England/QEUH-9B7C65/2020\|EPI_ISL_590519\|20 | ···················· | ···················· | ····················· | ···················· | ····T··············· | ···················· | |
| M | 7615 | hCoV-19/Australia/VIC11783/2020\|EPI_ISL_591791\|202 | ···················· | ···················· | ···K················· | ···················· | ···················· | ···················· | |
| M | 7616 | hCoV-19/Australia/VIC11804/2020\|EPI_ISL_591792\|202 | ···················· | ···················· | ····················· | ···················· | ···················· | ···················· | |
| M | 7617 | hCoV-19/Australia/VIC12942/2020\|EPI_ISL_591793\|202 | ···················· | ···················· | ····················· | ···················· | ···················· | ···················· | |
| M | 7619 | hCoV-19/Australia/VIC14134/2020\|EPI_ISL_591795\|202 | ···················· | ···················· | ····················· | ···················· | ···················· | ···················· | |
| M | 7620 | hCoV-19/Australia/VIC14341/2020\|EPI_ISL_591796\|202 | ···················· | ···················· | ····················· | ···················· | ···················· | ···················· | |
| M | 7621 | hCoV-19/Australia/VIC12846/2020\|EPI_ISL_591797\|202 | ···················· | ···················· | ····················· | ···················· | ···················· | ···················· | |
| M | 7622 | hCoV-19/Australia/VIC12859/2020\|EPI_ISL_591798\|202 | ···················· | ···················· | ····················· | ···················· | ···················· | ···················· | |
| M | 7623 | hCoV-19/Australia/VIC13084/2020\|EPI_ISL_591799\|202 | ···················· | ···················· | ···K················· | ···················· | ···················· | ···················· | |
| M | 7624 | hCoV-19/Australia/VIC13101/2020\|EPI_ISL_591800\|202 | ···················· | ···················· | ····················· | ···················· | ···················· | ···················· | |
| M | 7625 | hCoV-19/Australia/VIC13533/2020\|EPI_ISL_591801\|202 | ···················· | ···················· | ····················· | ···················· | ···················· | ···················· | |
| M | 7627 | hCoV-19/Australia/VIC14271/2020\|EPI_ISL_591803\|202 | ···················· | ···················· | ····················· | ···················· | ···················· | ···················· | |
| M | 7628 | hCoV-19/Australia/VIC14286/2020\|EPI_ISL_591804\|202 | ···················· | ···················· | ····················· | ···················· | ···················· | ···················· | |
| M | 7629 | hCoV-19/Australia/VIC14318/2020\|EPI_ISL_591805\|202 | ···················· | ···················· | ····················· | ···················· | ···················· | ···················· | |
| M | 7632 | hCoV-19/Australia/VIC11660/2020\|EPI_ISL_591830\|202 | ···················· | ···················· | ····················· | ···················· | ···················· | ···················· | |
| M | 7635 | hCoV-19/Australia/VIC11679/2020\|EPI_ISL_591833\|202 | ···················· | ···················· | ····················· | ···················· | ···················· | ···················· | |
| M | 7636 | hCoV-19/Australia/VIC11687/2020\|EPI_ISL_591834\|202 | ···················· | ···················· | ····················· | ···················· | ···················· | ···················· | |
| M | 7637 | hCoV-19/Australia/VIC11692/2020\|EPI_ISL_591835\|202 | ···················· | ···················· | ····················· | ···················· | ···················· | ···················· | |
| M | 7638 | hCoV-19/Australia/VIC11695/2020\|EPI_ISL_591836\|202 | ···················· | ···················· | ····················· | ···················· | ···················· | ···················· | |
| M | 7639 | hCoV-19/Australia/VIC11696/2020\|EPI_ISL_591837\|202 | ···················· | ···················· | ····················· | ···················· | ···················· | ···················· | |
| M | 7640 | hCoV-19/Australia/VIC11713/2020\|EPI_ISL_591838\|202 | ···················· | ···················· | ····················· | ···················· | ···················· | ···················· | |
| M | 7642 | hCoV-19/Australia/VIC11727/2020\|EPI_ISL_591840\|202 | ···················· | ···················· | ····················· | ···················· | ···················· | ···················· | |
| M | 7646 | hCoV-19/Australia/VIC11777/2020\|EPI_ISL_591848\|202 | ···················· | ···················· | ····················· | ···················· | NNN················· | ···················· | 1 |
| M | 7647 | hCoV-19/Australia/VIC11781/2020\|EPI_ISL_591849\|202 | ···················· | ···················· | ····················· | ···················· | ···················· | ···················· | |
| M | 7648 | hCoV-19/USA/UT-UPHL-201030072/2020\|EPI_ISL_594062\| | ···················· | ·A·················· | ····················· | ···················· | ···················· | ···················· | |
| M | 7649 | hCoV-19/USA/UT-UPHL-201019807/2020\|EPI_ISL_594063\| | ···················· | ···················· | ····················· | ···················· | ···················· | ···················· | |
| M | 7650 | hCoV-19/USA/UT-UPHL-201026584/2020\|EPI_ISL_594064\| | ···················· | ···················· | ····················· | ···················· | ···················· | ···················· | |
| M | 7651 | hCoV-19/USA/UT-UPHL-201011705/2020\|EPI_ISL_594066\| | ···················· | ···················· | ····················· | ···················· | ···················· | ···················· | |
| M | 7652 | hCoV-19/USA/UT-UPHL-201030137/2020\|EPI_ISL_594067\| | ···················· | ···················· | ····················· | ···················· | ···················· | ···················· | |
| M | 7653 | hCoV-19/USA/UT-UPHL-201025899/2020\|EPI_ISL_594069\| | ···················· | ···················· | ····················· | ···················· | ···················· | ···················· | |
| M | 7654 | hCoV-19/USA/UT-UPHL-201030995/2020\|EPI_ISL_594070\| | ···················· | ···················· | ····················· | ···················· | ···················· | ···················· | |
| M | 7655 | hCoV-19/USA/UT-UPHL-201021928/2020\|EPI_ISL_594071\| | ···················· | ···················· | ····················· | ···················· | ···················· | ···················· | |
| M | 7656 | hCoV-19/USA/UT-UPHL-201030943/2020\|EPI_ISL_594072\| | ···················· | ···················· | ····················· | ···················· | ···················· | ···················· | |
| M | 7657 | hCoV-19/USA/UT-UPHL-201009431/2020\|EPI_ISL_594074\| | ···················· | ···················· | ····················· | ···················· | ···················· | ···················· | |
| M | 7658 | hCoV-19/USA/UT-UPHL-201027345/2020\|EPI_ISL_594079\| | ···················· | ···················· | ····················· | ···················· | ···················· | ···················· | |
| M | 7659 | hCoV-19/USA/UT-UPHL-201022755/2020\|EPI_ISL_594081\| | ···················· | ···················· | ····················· | ···················· | ···················· | ···················· | |
| M | 7660 | hCoV-19/USA/UT-UPHL-201020113/2020\|EPI_ISL_594083\| | ···················· | ···················· | ····················· | ···················· | ···················· | ···················· | |
| M | 7661 | hCoV-19/USA/UT-UPHL-201029534/2020\|EPI_ISL_594084\| | ···················· | ···················· | ····················· | ···················· | ···················· | ···················· | |
| M | 7662 | hCoV-19/USA/UT-UPHL-201013892/2020\|EPI_ISL_594086\| | ···················· | ···················· | ····················· | ···················· | ···················· | ···················· | |
| M | 7663 | hCoV-19/Wales/PHWC-47DED3/2020\|EPI_ISL_595377\|2020 | ···················· | ···················· | ····················· | ···················· | ···················· | ···················· | |
| M | 7664 | hCoV-19/Wales/PHWC-47DEE2/2020\|EPI_ISL_595378\|2020 | ···················· | ···················· | ····················· | ···················· | ···················· | ···················· | |
| M | 7665 | hCoV-19/Wales/PHWC-47DF1C/2020\|EPI_ISL_595380\|2020 | ···················· | ···················· | ····················· | ···················· | ···················· | ···················· | |
| M | 7666 | hCoV-19/Wales/PHWC-47DF2B/2020\|EPI_ISL_595381\|2020 | ···················· | ···················· | ····················· | ···················· | ···················· | ···················· | |
| M | 7667 | hCoV-19/Wales/PHWC-47DF3A/2020\|EPI_ISL_595382\|2020 | ···················· | ···················· | ····················· | ···················· | ···················· | ···················· | |
| M | 7668 | hCoV-19/Wales/PHWC-47DF49/2020\|EPI_ISL_595383\|2020 | ···················· | ···················· | ····················· | ···················· | ···················· | ···················· | |
| M | 7669 | hCoV-19/Wales/PHWC-47DF76/2020\|EPI_ISL_595386\|2020 | ···················· | ···················· | ····················· | ···················· | ···················· | ···················· | |
| M | 7670 | hCoV-19/Wales/PHWC-47DF94/2020\|EPI_ISL_595388\|2020 | ···················· | ···················· | ····················· | ···················· | ···················· | ···················· | |
| M | 7671 | hCoV-19/Wales/PHWC-47DFA3/2020\|EPI_ISL_595389\|2020 | ···················· | ···················· | ····················· | ···················· | ···················· | ···················· | |
| M | 7672 | hCoV-19/Wales/PHWC-47DFB2/2020\|EPI_ISL_595390\|2020 | ···················· | ···················· | ····················· | ···················· | ···················· | ···················· | |
| M | 7673 | hCoV-19/Wales/PHWC-47DFC1/2020\|EPI_ISL_595391\|2020 | ···················· | ···················· | ····················· | ···················· | ···················· | ···················· | |
| M | 7674 | hCoV-19/Wales/PHWC-47DFD0/2020\|EPI_ISL_595392\|2020 | ···················· | ···················· | ····················· | ···················· | ···················· | ···················· | |
| M | 7675 | hCoV-19/Wales/PHWC-47DFEF/2020\|EPI_ISL_595393\|2020 | ···················· | ···················· | ····················· | ···················· | ···················· | ···················· | |
| M | 7676 | hCoV-19/Wales/PHWC-47DFFE/2020\|EPI_ISL_595394\|2020 | ···················· | ···················· | ····················· | ···················· | ···················· | ···················· | |
| M | 7677 | hCoV-19/Wales/PHWC-47E036/2020\|EPI_ISL_595396\|2020 | ···················· | ···················· | ····················· | ···················· | ···················· | ···················· | |
| M | 7678 | hCoV-19/Wales/PHWC-47E054/2020\|EPI_ISL_595397\|2020 | ···················· | ···················· | ····················· | ···················· | ···················· | ···················· | |
| M | 7679 | hCoV-19/Wales/PHWC-47E063/2020\|EPI_ISL_595398\|2020 | ···················· | ···················· | ····················· | ···················· | ···················· | ···················· | |
| M | 7681 | hCoV-19/England/LOND-124DB2F/2020\|EPI_ISL_584642\|2 | ···················· | ···················· | ····················· | ···················· | ···················· | ···················· | |
| M | 7682 | hCoV-19/England/LOND-124DB4D/2020\|EPI_ISL_584643\|2 | ···················· | ···················· | ····················· | ···················· | ···················· | ···················· | |
| M | 7683 | hCoV-19/England/LOND-124DC68/2020\|EPI_ISL_584645\|2 | ···················· | ···················· | ····················· | ···················· | ···················· | ···················· | |
| M | 7684 | hCoV-19/England/LOND-124DCE0/2020\|EPI_ISL_584646\|2 | ···················· | ···················· | ····················· | ···················· | ···················· | ···················· | |
| M | 7685 | hCoV-19/England/LOND-124DD29/2020\|EPI_ISL_584647\|2 | ···················· | ···················· | ····················· | ···················· | ···················· | ···················· | |
| M | 7688 | hCoV-19/England/LOND-124DE26/2020\|EPI_ISL_584651\|2 | ···················· | ···················· | ····················· | ···················· | ···················· | ···················· | |
| M | 7690 | hCoV-19/England/LOND-124DE62/2020\|EPI_ISL_584653\|2 | ···················· | ···················· | ····················· | ···················· | ···················· | ···················· | |
| M | 7691 | hCoV-19/England/LOND-124DF8D/2020\|EPI_ISL_584661\|2 | ···T················ | ···················· | ····················· | ···················· | ···················· | ···················· | |
| M | 7692 | hCoV-19/England/LOND-124E159/2020\|EPI_ISL_584662\|2 | ···················· | ·A·················· | ····················· | ···················· | ···················· | ···················· | |
| M | 7693 | hCoV-19/England/LOND-124E168/2020\|EPI_ISL_584663\|2 | ···················· | ···················· | ····················· | ···················· | ···················· | ···················· | |
| M | 7694 | hCoV-19/England/LOND-124E195/2020\|EPI_ISL_584664\|2 | ···················· | ···················· | ····················· | ···················· | ···················· | ···················· | |
| M | 7695 | hCoV-19/England/LOND-124E1B3/2020\|EPI_ISL_584666\|2 | ···················· | ···················· | ····················· | ···················· | ···················· | ···················· | |
| M | 7696 | hCoV-19/England/LOND-124E1FF/2020\|EPI_ISL_584668\|2 | ···················· | ···················· | ····················· | ···················· | ···················· | ···················· | |
| M | 7697 | hCoV-19/England/LOND-D9A6C/2020\|EPI_ISL_584674\|202 | ···················· | ·A·················· | ····················· | ···················· | ···················· | ···A················ | |
| M | 7698 | hCoV-19/England/LOND-D9A7B/2020\|EPI_ISL_584675\|202 | ···················· | ·A·················· | ····················· | ···················· | ···················· | ···A················ | |
| M | 7699 | hCoV-19/England/LOND-D9A8A/2020\|EPI_ISL_584676\|202 | ···················· | ···················· | ····················· | ···················· | ···················· | ···················· | |
| M | 7700 | hCoV-19/England/LOND-D9A99/2020\|EPI_ISL_584677\|202 | ···················· | ···················· | ····················· | ···················· | ···················· | ···················· | |
| M | 7701 | hCoV-19/England/LOND-D9AB7/2020\|EPI_ISL_584678\|202 | ···················· | ···················· | ····················· | ···················· | ···················· | ···················· | |
| M | 7702 | hCoV-19/England/LOND-D9AC6/2020\|EPI_ISL_584679\|202 | ···················· | ···················· | ····················· | ···················· | ···················· | ···················· | |
| M | 7703 | hCoV-19/England/MILK-9E1707/2020\|EPI_ISL_584680\|20 | ···················· | ···················· | ····················· | ···················· | ···················· | ···················· | |
| M | 7705 | hCoV-19/England/NORT-2A4F87/2020\|EPI_ISL_584682\|20 | ···················· | ···················· | ····················· | ···················· | ···················· | ···················· | |
| M | 7706 | hCoV-19/England/NORT-2A5065/2020\|EPI_ISL_584683\|20 | ···················· | ···················· | ····················· | ···················· | ···················· | ···················· | |

| | | | | | | | |
|---|---|---|---|---|---|---|---|
| M | 7707 hCoV-19/England/NORT-2A50DE/2020\|EPI_ISL_584684\|20 | ···················· | ···················· | ···················· | ···················· | ···················· | ···················· |
| M | 7708 hCoV-19/England/NORT-2A519F/2020\|EPI_ISL_584685\|20 | ···················· | ·A·················· | ···················· | ···················· | ···················· | ···················· |
| M | 7709 hCoV-19/England/NORT-2A5232/2020\|EPI_ISL_584686\|20 | ···················· | ···················· | ···················· | ···················· | ···················· | ···················· |
| M | 7710 hCoV-19/England/NORT-2A534E/2020\|EPI_ISL_584687\|20 | ···················· | ···················· | ···················· | ···················· | ···················· | ···················· |
| M | 7711 hCoV-19/England/NORT-2A5399/2020\|EPI_ISL_584688\|20 | ···················· | ···················· | ···················· | ···················· | ···················· | ···················· |
| M | 7712 hCoV-19/England/NORT-2A53F3/2020\|EPI_ISL_584689\|20 | ···················· | ···················· | ···················· | ···················· | ···················· | ···················· |
| M | 7713 hCoV-19/England/NORT-2A54D2/2020\|EPI_ISL_584690\|20 | ···················· | ···················· | ···················· | ···················· | ···················· | ···················· |
| M | 7714 hCoV-19/England/NORT-2A54E1/2020\|EPI_ISL_584691\|20 | ···················· | ···················· | ···················· | ···················· | ···················· | ···················· |
| M | 7715 hCoV-19/England/NORT-2A552A/2020\|EPI_ISL_584693\|20 | ···················· | ·A·················· | ···················· | ···················· | ···················· | ···················· |
| M | 7716 hCoV-19/England/NORT-2A55C0/2020\|EPI_ISL_584694\|20 | ···················· | ···················· | ···················· | ···················· | ···················· | ···················· |
| M | 7717 hCoV-19/England/NORT-2A5733/2020\|EPI_ISL_584696\|20 | ···················· | ·A·················· | ···················· | ···················· | ···················· | ···················· |
| M | 7718 hCoV-19/England/NORT-2A577F/2020\|EPI_ISL_584697\|20 | ···················· | ···················· | ···················· | ···················· | ···················· | ···················· |
| M | 7719 hCoV-19/England/NORT-2A578E/2020\|EPI_ISL_584698\|20 | ···················· | ···················· | ···················· | ···················· | ···················· | ···················· |
| M | 7720 hCoV-19/England/NORT-2A57AC/2020\|EPI_ISL_584699\|20 | ···················· | ···················· | ···················· | ···················· | ···················· | ···················· |
| M | 7721 hCoV-19/Wales/PHWC-16E9C8/2020\|EPI_ISL_585801\|2020 | ···················· | ···················· | ···················· | ···················· | ···················· | ···················· |
| M | 7722 hCoV-19/Wales/PHWC-16E9D7/2020\|EPI_ISL_585802\|2020 | ···················· | ···················· | ···················· | ···················· | ···················· | ···················· |
| M | 7723 hCoV-19/Wales/PHWC-16E9E6/2020\|EPI_ISL_585803\|2020 | ···················· | ···················· | ···················· | ···················· | ···················· | ···················· |
| M | 7724 hCoV-19/Wales/PHWC-16E9F5/2020\|EPI_ISL_585804\|2020 | ···················· | ···················· | ···················· | ···················· | ···················· | ···················· |
| M | 7725 hCoV-19/Wales/PHWC-16EA01/2020\|EPI_ISL_585805\|2020 | ···················· | ···················· | ···················· | ···················· | ···················· | ···················· |
| M | 7726 hCoV-19/Wales/PHWC-16EA10/2020\|EPI_ISL_585806\|2020 | ···················· | ···················· | ···················· | ···················· | ···················· | ···················· |
| M | 7728 hCoV-19/Wales/PHWC-16EA3E/2020\|EPI_ISL_585808\|2020 | ···················· | ···················· | ···················· | ···················· | ···················· | ···················· |
| M | 7729 hCoV-19/Wales/PHWC-16EA4D/2020\|EPI_ISL_585809\|2020 | ···················· | ···················· | ···················· | ···················· | ···················· | ···················· |
| M | 7730 hCoV-19/Wales/PHWC-16F4A8/2020\|EPI_ISL_585972\|2020 | ···················· | ···················· | ···················· | ···················· | ···················· | ···················· |
| M | 7732 hCoV-19/Wales/PHWC-16F4C6/2020\|EPI_ISL_585974\|2020 | ···················· | ···················· | ···················· | ···················· | ···················· | ···················· |
| M | 7733 hCoV-19/Wales/PHWC-16F4D5/2020\|EPI_ISL_585975\|2020 | ···················· | ···················· | ···················· | ···················· | ···················· | ···················· |
| M | 7734 hCoV-19/Wales/PHWC-16F4E4/2020\|EPI_ISL_585976\|2020 | ···················· | ···················· | ···················· | ···················· | ···················· | ···················· |
| M | 7736 hCoV-19/Wales/PHWC-16F50F/2020\|EPI_ISL_585978\|2020 | ···················· | ···················· | ···················· | ···················· | ···················· | ···················· |
| M | 7737 hCoV-19/Wales/PHWC-16F51E/2020\|EPI_ISL_585979\|2020 | ···················· | ···················· | ···················· | ···················· | ···················· | ···················· |
| M | 7738 hCoV-19/Wales/PHWC-16F52D/2020\|EPI_ISL_585980\|2020 | ···················· | ···················· | ···················· | ···················· | ···················· | ···················· |
| M | 7739 hCoV-19/Wales/PHWC-16F53C/2020\|EPI_ISL_585981\|2020 | ···················· | ···················· | ···················· | ···················· | ···················· | ···················· |
| M | 7740 hCoV-19/Wales/PHWC-16F54B/2020\|EPI_ISL_585982\|2020 | ···················· | ···················· | ···················· | ···················· | ···················· | ···················· |
| M | 7741 hCoV-19/Wales/PHWC-16F55A/2020\|EPI_ISL_585983\|2020 | ···················· | ···················· | ···················· | ···················· | ···················· | ···················· |
| M | 7742 hCoV-19/Wales/PHWC-16F569/2020\|EPI_ISL_585984\|2020 | ···················· | ···················· | ···················· | ···················· | ···················· | ···················· |
| M | 7743 hCoV-19/Wales/PHWC-16F578/2020\|EPI_ISL_585985\|2020 | ···················· | ···················· | ···················· | ···················· | ···················· | ···················· |
| M | 7744 hCoV-19/Wales/PHWC-16F587/2020\|EPI_ISL_585986\|2020 | ···················· | ···················· | ···················· | ···················· | ···················· | ···················· |
| M | 7745 hCoV-19/Wales/PHWC-16F596/2020\|EPI_ISL_585987\|2020 | ···················· | ···················· | ···················· | ···················· | ···················· | ···················· |
| M | 7746 hCoV-19/Wales/PHWC-16F5A5/2020\|EPI_ISL_585988\|2020 | ···················· | ···················· | ···················· | ···················· | ···················· | ···················· |
| M | 7747 hCoV-19/Wales/PHWC-16F5B4/2020\|EPI_ISL_585989\|2020 | ···················· | ···················· | ···················· | ···················· | ···················· | ···················· |
| M | 7748 hCoV-19/Wales/PHWC-16F5C3/2020\|EPI_ISL_585990\|2020 | ···················· | ···················· | ···················· | ···················· | ···················· | ···················· |
| M | 7749 hCoV-19/Wales/PHWC-16F5D2/2020\|EPI_ISL_585991\|2020 | ···················· | ···················· | ···················· | ···················· | ···················· | ···················· |
| M | 7750 hCoV-19/Wales/PHWC-16F5E1/2020\|EPI_ISL_585992\|2020 | ···················· | ···················· | ···················· | ···················· | ···················· | ···················· |
| M | 7751 hCoV-19/Wales/PHWC-16F5F0/2020\|EPI_ISL_585993\|2020 | ···················· | ···················· | ···················· | ···················· | ···················· | ···················· |
| M | 7752 hCoV-19/Wales/PHWC-16F61B/2020\|EPI_ISL_585995\|2020 | ···················· | ···················· | ···················· | ···················· | ···················· | ···················· |
| M | 7753 hCoV-19/Wales/PHWC-16F62A/2020\|EPI_ISL_585996\|2020 | ···················· | ···················· | ···················· | ···················· | ···················· | ···················· |
| M | 7754 hCoV-19/Wales/PHWC-16F639/2020\|EPI_ISL_585997\|2020 | ···················· | ···················· | ···················· | ···················· | ···················· | ···················· |
| M | 7755 hCoV-19/Wales/PHWC-16F648/2020\|EPI_ISL_585998\|2020 | ···················· | ···················· | ···················· | ···················· | ···················· | ···················· |
| M | 7756 hCoV-19/Wales/PHWC-16F657/2020\|EPI_ISL_585999\|2020 | ···················· | ···················· | ···················· | ···················· | ···················· | ···················· |
| M | 7757 hCoV-19/England/ALDP-9FD586/2020\|EPI_ISL_588051\|20 | ···················· | ···················· | ···················· | ···················· | ···················· | ···················· |
| M | 7758 hCoV-19/England/ALDP-9FD1DD/2020\|EPI_ISL_588052\|20 | ···················· | ···················· | ···················· | ···················· | ···················· | ···················· |
| M | 7759 hCoV-19/England/ALDP-9FD0EF/2020\|EPI_ISL_588053\|20 | ···················· | ···················· | ···················· | ···················· | ···················· | ···················· |
| M | 7760 hCoV-19/England/ALDP-9FCF98/2020\|EPI_ISL_588054\|20 | ···················· | ···················· | ···················· | ···················· | ···················· | ···················· |
| M | 7761 hCoV-19/England/ALDP-9FDE12/2020\|EPI_ISL_588055\|20 | ···················· | ···················· | ···················· | ···················· | ···················· | ···················· |
| M | 7762 hCoV-19/England/ALDP-9FDA1E/2020\|EPI_ISL_588058\|20 | ···················· | ·A·················· | ···················· | ···················· | ···················· | ···················· |
| M | 7763 hCoV-19/England/QEUH-9FAD45/2020\|EPI_ISL_588070\|20 | ···················· | ···················· | ···················· | ···················· | ···················· | ···················· |
| M | 7764 hCoV-19/England/QEUH-9FA765/2020\|EPI_ISL_588071\|20 | ···················· | ···················· | ···················· | ···················· | ····T··············· | ···················· |
| M | 7765 hCoV-19/England/QEUH-9FAE33/2020\|EPI_ISL_588072\|20 | ···················· | ···················· | ···················· | ···················· | ···················· | ···················· |
| M | 7766 hCoV-19/England/QEUH-9FAB69/2020\|EPI_ISL_588073\|20 | ···················· | ···················· | ···················· | ···················· | ······T············· | ···················· |
| M | 7767 hCoV-19/England/QEUH-9FADEB/2020\|EPI_ISL_588074\|20 | ········T··········· | ···················· | ···················· | ···················· | ···················· | ···················· |
| M | 7768 hCoV-19/England/QEUH-9FA686/2020\|EPI_ISL_588076\|20 | ···················· | ···················· | ···················· | ···················· | ···················· | ···················· |
| M | 7769 hCoV-19/England/QEUH-9FB649/2020\|EPI_ISL_588077\|20 | ···················· | ···················· | ···················· | ···················· | ···················· | ···················· |
| M | 7770 hCoV-19/England/QEUH-9FA4E6/2020\|EPI_ISL_588078\|20 | ···················· | ·A·················· | ···················· | ···················· | ···················· | ···················· |
| M | 7771 hCoV-19/England/QEUH-9FA5F2/2020\|EPI_ISL_588079\|20 | ···················· | ···················· | ···················· | ···················· | ···················· | ···················· |
| M | 7772 hCoV-19/England/QEUH-9FA871/2020\|EPI_ISL_588080\|20 | ···················· | ···················· | ···················· | ···················· | ···················· | ···················· |
| M | 7773 hCoV-19/England/QEUH-9FAB1E/2020\|EPI_ISL_588081\|20 | ···················· | ···················· | ···················· | ···················· | ····T··············· | ···················· |
| M | 7774 hCoV-19/England/QEUH-9FAA3F/2020\|EPI_ISL_588082\|20 | ···················· | ···················· | ···················· | ···················· | ···················· | ···················· |
| M | 7775 hCoV-19/England/QEUH-9FB227/2020\|EPI_ISL_588083\|20 | ···················· | ···················· | ···················· | ···················· | ···················· | ···················· |
| M | 7776 hCoV-19/England/QEUH-9FA5C5/2020\|EPI_ISL_588084\|20 | ···················· | ···················· | ···················· | ···················· | ···················· | ···················· |
| M | 7777 hCoV-19/England/QEUH-9FACC0/2020\|EPI_ISL_588085\|20 | ···················· | ·A·················· | ···················· | ···················· | ···················· | ···················· |
| M | 7778 hCoV-19/England/QEUH-9FA6A4/2020\|EPI_ISL_588086\|20 | ···················· | ···················· | ···················· | ···················· | ···················· | ···················· |
| M | 7779 hCoV-19/England/QEUH-9FA6D1/2020\|EPI_ISL_588088\|20 | ···················· | ···················· | ···················· | ···················· | ···················· | ···················· |
| M | 7780 hCoV-19/England/QEUH-9FB263/2020\|EPI_ISL_588089\|20 | ···················· | ···················· | ···················· | ···················· | ···················· | ···················· |
| M | 7781 hCoV-19/England/ALDP-9ED3E9/2020\|EPI_ISL_589380\|20 | ···················· | ···················· | ···················· | ···················· | ···················· | ···················· |
| M | 7782 hCoV-19/England/ALDP-9E5967/2020\|EPI_ISL_589381\|20 | ···················· | ···················· | ···················· | ···················· | ···················· | ···················· |
| M | 7783 hCoV-19/England/ALDP-9ED2FB/2020\|EPI_ISL_589382\|20 | ···················· | ···················· | ···················· | ···················· | ···················· | ···················· |
| M | 7784 hCoV-19/England/ALDP-9ED4B9/2020\|EPI_ISL_589383\|20 | ···················· | ···················· | ···················· | ···················· | ···················· | ···················· |
| M | 7785 hCoV-19/England/ALDP-9EE078/2020\|EPI_ISL_589384\|20 | ···················· | ···················· | ···················· | ···················· | ···················· | ···················· |
| M | 7786 hCoV-19/England/ALDP-9E584C/2020\|EPI_ISL_589385\|20 | ···················· | ···················· | ···················· | ···················· | ···················· | ···················· |
| M | 7787 hCoV-19/England/ALDP-9ED0A6/2020\|EPI_ISL_589386\|20 | ···················· | ···················· | ···················· | ···················· | ···················· | ···················· |
| M | 7788 hCoV-19/England/ALDP-9ED149/2020\|EPI_ISL_589387\|20 | ···················· | ···················· | ···················· | ···················· | ···················· | ···················· |
| M | 7789 hCoV-19/England/ALDP-9ECF7D/2020\|EPI_ISL_589388\|20 | ···················· | ···················· | ···················· | ···················· | ···················· | ···················· |

| | | | | | | | | |
|---|---|---|---|---|---|---|---|---|
| M | 7790 hCoV-19/England/ALDP-9EDF7C/2020\|EPI_ISL_589389\|20 | ···················· | ···················· | ····················· | ···················· | ···················· | ···················· | |
| M | 7791 hCoV-19/England/ALDP-9EE03C/2020\|EPI_ISL_589399\|20 | ···················· | ···················· | ····················· | ···················· | ···················· | ···················· | |
| M | 7792 hCoV-19/England/ALDP-9ED62C/2020\|EPI_ISL_589500\|20 | ···················· | ···················· | ····················· | ···················· | ···················· | ···················· | |
| M | 7793 hCoV-19/England/ALDP-9ED3AD/2020\|EPI_ISL_589501\|20 | ···················· | ···················· | ····················· | ···················· | ···················· | ···················· | |
| M | 7794 hCoV-19/England/ALDP-9ED6B3/2020\|EPI_ISL_589502\|20 | ···················· | ···················· | ····················· | ···················· | ···················· | ···················· | |
| M | 7795 hCoV-19/England/ALDP-9E577C/2020\|EPI_ISL_589503\|20 | ···················· | ···················· | ····················· | ···················· | ···················· | ···················· | |
| M | 7796 hCoV-19/England/ALDP-9ED05B/2020\|EPI_ISL_589504\|20 | ···················· | ···················· | ····················· | ···················· | ···················· | ···················· | |
| M | 7797 hCoV-19/England/ALDP-9E5CD7/2020\|EPI_ISL_589505\|20 | ···················· | ···················· | ····················· | ···················· | ···················· | ···················· | |
| M | 7798 hCoV-19/England/ALDP-9ED097/2020\|EPI_ISL_589506\|20 | ···················· | ···················· | ····················· | ···················· | ···················· | ···················· | |
| M | 7799 hCoV-19/England/ALDP-9ED316/2020\|EPI_ISL_589507\|20 | ···················· | ···················· | ····················· | ···················· | ···················· | ···················· | |
| M | 7800 hCoV-19/England/ALDP-9E58D3/2020\|EPI_ISL_589508\|20 | ···················· | ···················· | ····················· | ···················· | ···················· | ···················· | |
| M | 7801 hCoV-19/England/ALDP-9E58C4/2020\|EPI_ISL_589509\|20 | ···················· | ···················· | ····················· | ···················· | ···················· | ···················· | |
| M | 7802 hCoV-19/England/ALDP-9ED000/2020\|EPI_ISL_589510\|20 | ···················· | ···················· | ····················· | ···················· | ···················· | ···················· | |
| M | 7803 hCoV-19/England/ALDP-9E5A64/2020\|EPI_ISL_589511\|20 | ···················· | ···················· | ····················· | ···················· | ···················· | ···················· | |
| M | 7804 hCoV-19/England/ALDP-9ED46E/2020\|EPI_ISL_589512\|20 | ···················· | ···················· | ····················· | ···················· | ···················· | ···················· | |
| M | 7805 hCoV-19/England/ALDP-9E5C13/2020\|EPI_ISL_589513\|20 | ···················· | ···················· | ····················· | ···················· | ···················· | ···················· | |
| M | 7806 hCoV-19/England/ALDP-9B5B79/2020\|EPI_ISL_590361\|20 | ···················· | ···················· | ········T············ | ···················· | ·····T·············· | ···················· | |
| M | 7807 hCoV-19/England/QEUH-9B4D65/2020\|EPI_ISL_590362\|20 | ···················· | ···················· | ····················· | ···················· | ···················· | ···················· | |
| M | 7808 hCoV-19/England/QEUH-9B506B/2020\|EPI_ISL_590363\|20 | ···················· | ···················· | ····················· | ···················· | ···················· | ···················· | |
| M | 7809 hCoV-19/England/ALDP-9B5906/2020\|EPI_ISL_590364\|20 | ···················· | ···················· | ····················· | ···················· | ···················· | ···················· | |
| M | 7810 hCoV-19/England/QEUH-9B4DFC/2020\|EPI_ISL_590365\|20 | ···················· | ·A·················· | ····················· | ···················· | ···················· | ··A················· | |
| M | 7811 hCoV-19/England/ALDP-9B5784/2020\|EPI_ISL_590366\|20 | ···················· | ···················· | ····················· | ···················· | ···················· | ···················· | |
| M | 7812 hCoV-19/England/ALDP-9B57A2/2020\|EPI_ISL_590367\|20 | ···················· | ···················· | ····················· | ···················· | ···················· | ···················· | |
| M | 7813 hCoV-19/England/QEUH-9B4DED/2020\|EPI_ISL_590368\|20 | ···················· | ···················· | ····················· | ···················· | ···················· | ···················· | |
| M | 7814 hCoV-19/England/ALDP-9B56E1/2020\|EPI_ISL_590369\|20 | ···················· | ···················· | ····················· | ···················· | ···················· | ···················· | |
| M | 7815 hCoV-19/England/QEUH-9B4BE3/2020\|EPI_ISL_590370\|20 | ···················· | ···················· | ····················· | ···················· | ···················· | ···················· | |
| M | 7816 hCoV-19/England/QEUH-9B4D56/2020\|EPI_ISL_590371\|20 | ···················· | ···················· | ····················· | ···················· | ···················· | ···················· | |
| M | 7817 hCoV-19/England/QEUH-9B4864/2020\|EPI_ISL_590376\|20 | ···················· | ···················· | ····················· | ···················· | ···················· | ···················· | |
| M | 7818 hCoV-19/England/QEUH-9B50C5/2020\|EPI_ISL_590377\|20 | ···················· | ···················· | ····················· | ···················· | ···················· | ···················· | |
| M | 7819 hCoV-19/England/ALDP-9B5BA6/2020\|EPI_ISL_590380\|20 | ···················· | ···················· | ········T············ | ···················· | ·····T·············· | ···················· | |
| M | 7820 hCoV-19/England/QEUH-9B5520/2020\|EPI_ISL_590381\|20 | ···················· | ···················· | ····················· | ···················· | ···················· | ···················· | |
| M | 7821 hCoV-19/England/QEUH-9B4943/2020\|EPI_ISL_590382\|20 | ···················· | ···················· | ····················· | ···················· | ···················· | ···················· | |
| M | 7822 hCoV-19/England/ALDP-9B5B5B/2020\|EPI_ISL_590383\|20 | ···················· | ···················· | ····················· | ···················· | ···················· | ···················· | |
| M | 7823 hCoV-19/England/QEUH-9B5696/2020\|EPI_ISL_590384\|20 | ···················· | ···················· | ····················· | ···················· | ···················· | ···················· | |
| M | 7824 hCoV-19/England/QEUH-9B49DA/2020\|EPI_ISL_590385\|20 | ···················· | ···················· | ····················· | ···················· | ···················· | ···················· | |
| M | 7825 hCoV-19/England/ALDP-9B5AC7/2020\|EPI_ISL_590386\|20 | ···················A | ···················· | ····················· | ···················· | ···················· | ···················· | 1 |
| M | 7826 hCoV-19/England/QEUH-9B51FF/2020\|EPI_ISL_590387\|20 | ···················· | ·A·················· | ····················· | ···················· | ···················· | ···················· | |
| M | 7827 hCoV-19/England/ALDP-9B5A21/2020\|EPI_ISL_590388\|20 | ···················· | ···················· | ····················· | ···················· | ···················· | ···················· | |
| M | 7828 hCoV-19/England/ALDP-9B5C0D/2020\|EPI_ISL_590390\|20 | ···················· | ···················· | ····················· | ···················· | ···················· | ···················· | |
| M | 7829 hCoV-19/England/QEUH-9B4F7E/2020\|EPI_ISL_590391\|20 | ···················· | ···················· | ····················· | ···················· | ···················· | ···················· | |
| M | 7830 hCoV-19/England/ALDP-9B5775/2020\|EPI_ISL_590392\|20 | ···················· | ···················· | ····················· | ···················· | ···················· | ···················· | |
| M | 7831 hCoV-19/England/QEUH-9B53F9/2020\|EPI_ISL_590393\|20 | ···················· | ···················· | ····················· | ···················· | ···················· | ···················· | |
| M | 7832 hCoV-19/England/QEUH-9B5687/2020\|EPI_ISL_590394\|20 | ···················· | ···················· | ····················· | ···················· | ···················· | ···················· | |
| M | 7833 hCoV-19/England/QEUH-9B47A3/2020\|EPI_ISL_590395\|20 | ···················· | ···················· | ····················· | ···················· | ···················· | ···················· | |
| M | 7834 hCoV-19/England/QEUH-9B4C95/2020\|EPI_ISL_590396\|20 | ···················· | ···················· | ····················· | ···················· | ···················· | ···············R···· | |
| M | 7835 hCoV-19/England/ALDP-9B5CEF/2020\|EPI_ISL_590397\|20 | ···················· | ···················· | ····················· | ···················· | ···················· | ···················· | |
| M | 7836 hCoV-19/England/ALDP-9B5B6A/2020\|EPI_ISL_590398\|20 | ···················· | ···················· | ········T············ | ···················· | ·····T·············· | ···················· | |
| M | 7837 hCoV-19/England/ALDP-9B5B88/2020\|EPI_ISL_590399\|20 | ···················· | ···················· | ····················· | ···················· | ···················· | ···················· | |
| M | 7838 hCoV-19/England/QEUH-9B703C/2020\|EPI_ISL_590512\|20 | ···················· | ·A·················· | ····················· | ···················· | ···················· | ··A················· | |
| M | 7839 hCoV-19/England/QEUH-9B7913/2020\|EPI_ISL_590514\|20 | ···················· | ···················· | ····················· | ···················· | ···················· | ···················· | |
| M | 7840 hCoV-19/England/QEUH-9B7500/2020\|EPI_ISL_590515\|20 | ···················· | ···················· | ····················· | ···················· | ···················· | ···················· | |
| M | 7842 hCoV-19/Australia/VIC14145/2020\|EPI_ISL_591819\|202 | ···················· | ···················· | ····················· | ···················· | N··················· | ···················· | |
| M | 7843 hCoV-19/Australia/VIC14148/2020\|EPI_ISL_591820\|202 | ···················· | ···················· | ····················· | ···················· | ···················· | ···················· | |
| M | 7844 hCoV-19/Australia/VIC14250/2020\|EPI_ISL_591821\|202 | ···················· | ···················· | ····················· | ···················· | ···················· | ···················· | |
| M | 7847 hCoV-19/Australia/VIC11630/2020\|EPI_ISL_591828\|202 | ···················· | ···················· | ····················· | ···················· | ···················· | ···················· | |
| M | 7849 hCoV-19/Australia/VIC11751/2020\|EPI_ISL_591843\|202 | ···················· | ···················· | ····················· | ···················· | ···················· | ···················· | |
| M | 7850 hCoV-19/Australia/VIC11755/2020\|EPI_ISL_591844\|202 | ···················· | ···················· | ····················· | ···················· | ···················· | ···················· | |
| M | 7851 hCoV-19/Australia/VIC11766/2020\|EPI_ISL_591845\|202 | ···················· | ···················· | ····················· | ···················· | ···················· | ···················· | |
| M | 7852 hCoV-19/Australia/VIC11775/2020\|EPI_ISL_591846\|202 | ···················· | ···················· | ····················· | ···················· | ···················· | ···················· | |
| M | 7853 hCoV-19/USA/UT-UPHL-201020175/2020\|EPI_ISL_594007\| | ···················· | ···················· | ····················· | ···················· | ···················· | ···················· | |
| M | 7854 hCoV-19/USA/UT-UPHL-201027142/2020\|EPI_ISL_594008\| | ···················· | ···················· | ····················· | ···················· | ···················· | ···················· | |
| M | 7855 hCoV-19/USA/UT-UPHL-201004118/2020\|EPI_ISL_594009\| | ···················· | ···················· | ····················· | ···················· | ···················· | ···················· | |
| M | 7856 hCoV-19/USA/UT-UPHL-201014531/2020\|EPI_ISL_594010\| | ···················· | ···················· | ····················· | ···················· | ···················· | ···················· | |
| M | 7857 hCoV-19/USA/UT-UPHL-201029634/2020\|EPI_ISL_594013\| | ···················· | ···················· | ····················· | ···················· | ···················· | ···················· | |
| M | 7858 hCoV-19/USA/UT-UPHL-201009890/2020\|EPI_ISL_594014\| | ···················· | ···················· | ····················· | ···················· | ···················· | ···················· | |
| M | 7859 hCoV-19/USA/UT-UPHL-201031247/2020\|EPI_ISL_594015\| | ···················· | ···················· | ····················· | ···················· | ···················· | ···················· | |
| M | 7860 hCoV-19/USA/UT-UPHL-201016782/2020\|EPI_ISL_594017\| | ···················· | ···················· | ····················· | ···················· | ···················· | ···················· | |
| M | 7861 hCoV-19/USA/UT-UPHL-201008114/2020\|EPI_ISL_594018\| | ···················· | ···················· | ····················· | ···················· | ···················· | ···················· | |
| M | 7862 hCoV-19/USA/UT-UPHL-201016722/2020\|EPI_ISL_594021\| | ···················· | ···················· | ····················· | ···················· | ···················· | ···················· | |
| M | 7863 hCoV-19/USA/UT-UPHL-201022870/2020\|EPI_ISL_594026\| | ···················· | ···················· | ····················· | ···················· | ···················· | ···················· | |
| M | 7864 hCoV-19/USA/UT-UPHL-201009152/2020\|EPI_ISL_594027\| | ···················· | ···················· | ····················· | ···················· | ···················· | ···················· | |
| M | 7865 hCoV-19/USA/UT-UPHL-201023823/2020\|EPI_ISL_594034\| | ···················· | ·A·················· | ····················· | ···················· | ···················· | ···················· | |
| M | 7866 hCoV-19/USA/UT-UPHL-201016479/2020\|EPI_ISL_594035\| | ···················· | ···················· | ····················· | ···················· | ···················· | ···················· | |
| M | 7867 hCoV-19/USA/UT-UPHL-201010816/2020\|EPI_ISL_594038\| | ···················· | ·A·················· | ····················· | ···················· | ···················· | ···················· | |
| M | 7868 hCoV-19/USA/UT-UPHL-201012960/2020\|EPI_ISL_594039\| | ···················· | ···················· | ····················· | ···················· | ···················· | ···················· | |
| M | 7869 hCoV-19/USA/UT-UPHL-201005514/2020\|EPI_ISL_594040\| | ···················· | ···················· | ····················· | ···················· | ···················· | ···················· | |
| M | 7870 hCoV-19/USA/UT-UPHL-201023541/2020\|EPI_ISL_594042\| | ···················· | ···················· | ····················· | ···················· | ···················· | ···················· | |
| M | 7871 hCoV-19/USA/UT-UPHL-201015242/2020\|EPI_ISL_594091\| | ···················· | ···················· | ····················· | ···················· | ···················· | ···················· | |
| M | 7872 hCoV-19/USA/UT-UPHL-201000140/2020\|EPI_ISL_594095\| | ···················· | ···················· | ····················· | ···················· | ···················· | ···················· | |
| M | 7873 hCoV-19/USA/UT-UPHL-201016311/2020\|EPI_ISL_594097\| | ···················· | ···················· | ····················· | ···················· | ···················· | ···················· | |

| | | | | | | | | |
|---|---|---|---|---|---|---|---|---|
| M | 7874 hCoV-19/USA/UT-UPHL-201032575/2020\|EPI_ISL_594098\| | ···················· | ···················· | ···················· | ···················· | ···················· | ···················· | |
| M | 7875 hCoV-19/USA/UT-UPHL-201020918/2020\|EPI_ISL_594099\| | ···················· | ···················· | ···················· | ···················· | ···················· | ···················· | |
| M | 7876 hCoV-19/England/NOTT-115FA0/2020\|EPI_ISL_595300\|20 | ···················· | ···················· | ···················· | ···················· | ···················· | ···················· | |
| M | 7877 hCoV-19/England/NOTT-116060/2020\|EPI_ISL_595302\|20 | ···················· | ···················· | ···················· | ···················· | ···················· | ···················· | |
| M | 7878 hCoV-19/England/NOTT-116112/2020\|EPI_ISL_595303\|20 | ···················· | ···················· | ···················· | ···················· | ···················· | ···················· | |
| M | 7879 hCoV-19/England/NOTT-116121/2020\|EPI_ISL_595304\|20 | ···················· | ···················· | ···················· | ···················· | ···················· | ···················· | |
| M | 7880 hCoV-19/England/NOTT-11614F/2020\|EPI_ISL_595305\|20 | ···················· | ···················· | ···················· | ···················· | ···················· | ···················· | |
| M | 7881 hCoV-19/England/NOTT-11616D/2020\|EPI_ISL_595306\|20 | ···················· | ···················· | ···················· | ···················· | ···················· | ···················· | |
| M | 7882 hCoV-19/England/NOTT-1161B8/2020\|EPI_ISL_595307\|20 | ···················· | ···················· | ···················· | ···················· | ···················· | ···················· | |
| M | 7883 hCoV-19/England/NOTT-1161D6/2020\|EPI_ISL_595308\|20 | ···················· | ···················· | ···················· | ···················· | ···················· | ···················· | |
| M | 7884 hCoV-19/England/NOTT-1161E5/2020\|EPI_ISL_595309\|20 | ···················· | ···················· | ···················· | ···················· | ···················· | ···················· | |
| M | 7885 hCoV-19/England/NOTT-1161F4/2020\|EPI_ISL_595310\|20 | ···················· | ···················· | ···················· | ···················· | ···················· | ···················· | |
| M | 7886 hCoV-19/England/NOTT-116200/2020\|EPI_ISL_595311\|20 | ···················· | ···················· | ···················· | ···················· | ···················· | ···················· | |
| M | 7887 hCoV-19/England/NOTT-11621F/2020\|EPI_ISL_595312\|20 | ···················· | ···················· | ···················· | ···················· | ···················· | ···················· | |
| M | 7888 hCoV-19/England/NOTT-116349/2020\|EPI_ISL_595315\|20 | ···················· | ···················· | ···················· | ···················· | ···················· | ···················· | |
| M | 7889 hCoV-19/England/NOTT-116798/2020\|EPI_ISL_595317\|20 | ···················· | ···················· | ···················· | ···················· | ···················· | ···················· | |
| M | 7890 hCoV-19/England/NOTT-1167A7/2020\|EPI_ISL_595318\|20 | ···················· | ···················· | ···················· | ···················· | ···················· | ···················· | |
| M | 7891 hCoV-19/England/NOTT-1167E3/2020\|EPI_ISL_595319\|20 | ···················· | ···················· | ···················· | ···················· | ···················· | ···················· | |
| M | 7892 hCoV-19/England/NOTT-116ADB/2020\|EPI_ISL_595320\|20 | ···················· | ···················· | ···················· | ···················· | ···················· | ···················· | |
| M | 7893 hCoV-19/England/NOTT-116B6F/2020\|EPI_ISL_595322\|20 | ···················· | ···················· | ···················· | ···················· | ···················· | ···················· | |
| M | 7894 hCoV-19/England/NOTT-116C11/2020\|EPI_ISL_595323\|20 | ···················· | ···················· | ···················· | ···················· | ···················· | ···················· | |
| M | 7895 hCoV-19/England/NOTT-116DE1/2020\|EPI_ISL_595324\|20 | ···················· | ···················· | ···················· | ···················· | ···················· | ···················· | |
| M | 7896 hCoV-19/England/NOTT-116EEE/2020\|EPI_ISL_595325\|20 | ···················· | ···················· | ···················· | ···················· | ···················· | ···················· | |
| M | 7897 hCoV-19/England/OXON-B518F/2020\|EPI_ISL_595326\|202 | ···················· | ···················· | ···················· | ···················· | ···················· | ···················· | |
| M | 7898 hCoV-19/England/OXON-B51CB/2020\|EPI_ISL_595327\|202 | ···················· | ···················· | ···················· | ···················· | ·····T·············· | ···················· | |
| M | 7899 hCoV-19/England/OXON-B51DA/2020\|EPI_ISL_595328\|202 | ···················· | ···················· | ···················· | ···················· | ···················· | ···················· | |
| M | 7900 hCoV-19/Wales/PHWC-47DB18/2020\|EPI_ISL_595329\|2020 | ···················· | ···················· | ···················· | ···················· | ···················· | ···················· | |
| M | 7901 hCoV-19/Wales/PHWC-47DB36/2020\|EPI_ISL_595330\|2020 | ···················· | ···················· | ···················· | ···················· | ···················· | ···················· | |
| M | 7902 hCoV-19/Wales/PHWC-47DB45/2020\|EPI_ISL_595331\|2020 | ···················· | ···················· | ···················· | ···················· | ···················· | ···················· | |
| M | 7903 hCoV-19/Wales/PHWC-47DB54/2020\|EPI_ISL_595332\|2020 | ···················· | ···················· | ···················· | ···················· | ···················· | ···················· | |
| M | 7904 hCoV-19/Wales/PHWC-47DB63/2020\|EPI_ISL_595333\|2020 | ···················· | ···················· | ···················· | ···················· | ···················· | ···················· | |
| M | 7906 hCoV-19/Wales/PHWC-47DBBE/2020\|EPI_ISL_595336\|2020 | ···················· | ···················· | ···················· | ···················· | ···················· | ···················· | |
| M | 7907 hCoV-19/Wales/PHWC-47DBCD/2020\|EPI_ISL_595337\|2020 | ···················· | ···················· | ···················· | ···················· | ···················· | ···················· | |
| M | 7908 hCoV-19/Wales/PHWC-47DBDC/2020\|EPI_ISL_595338\|2020 | ···················· | ···················· | ···················· | ···················· | NNN················· | ···················· | 1 |
| M | 7909 hCoV-19/Wales/PHWC-47DBEB/2020\|EPI_ISL_595339\|2020 | ···················· | ···················· | ···················· | ···················· | ···················· | ···················· | |
| M | 7910 hCoV-19/Wales/PHWC-47DBFA/2020\|EPI_ISL_595340\|2020 | ···················· | ···················· | ···················· | ···················· | ···················· | ···················· | |
| M | 7911 hCoV-19/Wales/PHWC-47DC06/2020\|EPI_ISL_595341\|2020 | ···················· | ···················· | ···················· | ···················· | ···················· | ···················· | |
| M | 7912 hCoV-19/Wales/PHWC-47DC15/2020\|EPI_ISL_595342\|2020 | ···················· | ···················· | ···················· | ···················· | ···················· | ···················· | |
| M | 7913 hCoV-19/Wales/PHWC-47DC24/2020\|EPI_ISL_595343\|2020 | ···················· | ···················· | ···················· | ···················· | ···················· | ···················· | |
| M | 7914 hCoV-19/Wales/PHWC-47DC51/2020\|EPI_ISL_595344\|2020 | ···················· | ···················· | ···················· | ···················· | ···················· | ···················· | |
| M | 7915 hCoV-19/Wales/PHWC-47DC7F/2020\|EPI_ISL_595346\|2020 | ···················· | ···················· | ···················· | ···················· | ···················· | ···················· | |
| M | 7917 hCoV-19/Wales/PHWC-47DCAC/2020\|EPI_ISL_595349\|2020 | ···················· | ···················· | ···················· | ···················· | ···················· | ···················· | |
| M | 7918 hCoV-19/Wales/PHWC-47DCBB/2020\|EPI_ISL_595350\|2020 | ···················· | ···················· | ···················· | ···················· | ···················· | ···················· | |
| M | 7920 hCoV-19/Wales/PHWC-47DCF7/2020\|EPI_ISL_595352\|2020 | ···················· | ···················· | ···················· | ···················· | ···················· | ···················· | |
| M | 7921 hCoV-19/Wales/PHWC-47DD21/2020\|EPI_ISL_595355\|2020 | ···················· | ···················· | ···················· | ···················· | ···················· | ···················· | |
| M | 7922 hCoV-19/Wales/PHWC-47DD4F/2020\|EPI_ISL_595357\|2020 | ···················· | ···················· | ···················· | ···················· | ···················· | ···················· | |
| M | 7923 hCoV-19/Wales/PHWC-47DD5E/2020\|EPI_ISL_595358\|2020 | ···················· | ···················· | ···················· | ···················· | ···················· | ···················· | |
| M | 7924 hCoV-19/Wales/PHWC-47DD6D/2020\|EPI_ISL_595359\|2020 | ···················· | ···················· | ···················· | ···················· | ···················· | ···················· | |
| M | 7926 hCoV-19/Wales/PHWC-47DDC7/2020\|EPI_ISL_595363\|2020 | ···················· | ···················· | ···················· | ···················· | ···················· | ···················· | |
| M | 7927 hCoV-19/Wales/PHWC-47DDD6/2020\|EPI_ISL_595364\|2020 | ···················· | ···················· | ···················· | ···················· | ···················· | ···················· | |
| M | 7928 hCoV-19/Wales/PHWC-47DDE5/2020\|EPI_ISL_595365\|2020 | ···················· | ···················· | ···················· | ···················· | ···················· | ···················· | |
| M | 7929 hCoV-19/Wales/PHWC-47DDF4/2020\|EPI_ISL_595366\|2020 | ···················· | ···················· | ···················· | ···················· | ··N················· | ···················· | |
| M | 7932 hCoV-19/Wales/PHWC-47DE4C/2020\|EPI_ISL_595369\|2020 | ···················· | ···················· | ···················· | ···················· | ···················· | ···················· | |
| M | 7933 hCoV-19/Wales/PHWC-47DE6A/2020\|EPI_ISL_595371\|2020 | ···················· | ···················· | ···················· | ···················· | ···················· | ···················· | |
| M | 7934 hCoV-19/Wales/PHWC-47DE79/2020\|EPI_ISL_595372\|2020 | ···················· | ···················· | ···················· | ···················· | ···················· | ···················· | |
| M | 7935 hCoV-19/Wales/PHWC-47DE97/2020\|EPI_ISL_595374\|2020 | ···················· | ···················· | ···················· | ···················· | ···················· | ···················· | |
| M | 7936 hCoV-19/Wales/PHWC-47DEC4/2020\|EPI_ISL_595376\|2020 | ···················· | ···················· | ···················· | ···················· | ···················· | ···················· | |
| M | 7937 hCoV-19/England/CWAR-24FDC44/2020\|EPI_ISL_584500\|2 | ···················· | ···················· | ···················· | ···················· | ···················· | ···················· | |
| M | 7938 hCoV-19/England/CWAR-24FDC80/2020\|EPI_ISL_584504\|2 | ···················· | ···················· | ···················· | ···················· | ···················· | ···················· | |
| M | 7939 hCoV-19/England/CWAR-24FDC9F/2020\|EPI_ISL_584505\|2 | ···················· | ···················· | ···················· | ···················· | ···················· | ···················· | |
| M | 7940 hCoV-19/England/CWAR-24FDCAE/2020\|EPI_ISL_584506\|2 | ···················· | ···················· | ···················· | ···················· | ···················· | ···················· | |
| M | 7941 hCoV-19/England/CWAR-24FDCBD/2020\|EPI_ISL_584507\|2 | ···················· | ···················· | ···················· | ···················· | ···················· | ···················· | |
| M | 7942 hCoV-19/England/CWAR-24FDCCC/2020\|EPI_ISL_584508\|2 | ···················· | ···················· | ···················· | ···················· | ···················· | ···················· | |
| M | 7943 hCoV-19/England/CWAR-24FDCEA/2020\|EPI_ISL_584510\|2 | ···················· | ···················· | ···················· | ···················· | ···················· | ···················· | |
| M | 7944 hCoV-19/England/CWAR-24FDCF9/2020\|EPI_ISL_584511\|2 | ···················· | ···················· | ···················· | ···················· | ···················· | ···················· | |
| M | 7945 hCoV-19/England/CWAR-24FDD05/2020\|EPI_ISL_584512\|2 | ···················· | ···················· | ···················· | ···················· | ···················· | ···················· | |
| M | 7946 hCoV-19/England/CWAR-24FDD14/2020\|EPI_ISL_584513\|2 | ···················· | ···················· | ···················· | ···················· | ···················· | ···················· | |
| M | 7947 hCoV-19/England/CWAR-24FDD23/2020\|EPI_ISL_584514\|2 | ···················· | ···················· | ···················· | ···················· | ···················· | ···················· | |
| M | 7948 hCoV-19/England/CWAR-24FDD32/2020\|EPI_ISL_584515\|2 | ···················· | ···················· | ···················· | ···················· | ···················· | ···················· | |
| M | 7949 hCoV-19/England/CWAR-24FDD41/2020\|EPI_ISL_584516\|2 | ···················· | ···················· | ···················· | ···················· | ···················· | ···················· | |
| M | 7950 hCoV-19/England/CWAR-24FDD50/2020\|EPI_ISL_584517\|2 | ···················· | ···················· | ···················· | ···················· | ···················· | ···················· | |
| M | 7951 hCoV-19/England/CWAR-24FDD6F/2020\|EPI_ISL_584518\|2 | ···················· | ···················· | ···················· | ···················· | ···················· | ···················· | |
| M | 7952 hCoV-19/England/CWAR-24FDD7E/2020\|EPI_ISL_584519\|2 | ···················· | ···················· | ···················· | ···················· | ···················· | ···················· | |
| M | 7953 hCoV-19/Wales/PHWC-16EA5C/2020\|EPI_ISL_585810\|2020 | ···················· | ···················· | ···················· | ···················· | ···················· | ···················· | |
| M | 7954 hCoV-19/Wales/PHWC-16EA6B/2020\|EPI_ISL_585811\|2020 | ···················· | ···················· | ···················· | ···················· | ···················· | ···················· | |
| M | 7955 hCoV-19/Wales/PHWC-16EA7A/2020\|EPI_ISL_585812\|2020 | ···················· | ···················· | ···················· | ···················· | ···················· | ···················· | |
| M | 7956 hCoV-19/Wales/PHWC-16EA89/2020\|EPI_ISL_585813\|2020 | ···················· | ···················· | ···················· | ···················· | ···················· | ···················· | |
| M | 7957 hCoV-19/Wales/PHWC-16EA98/2020\|EPI_ISL_585814\|2020 | ···················· | ···················· | ···················· | ···················· | ···················· | ···················· | |
| M | 7958 hCoV-19/Wales/PHWC-16EAA7/2020\|EPI_ISL_585815\|2020 | ···················· | ···················· | ···················· | ···················· | ···················· | ···················· | |
| M | 7959 hCoV-19/Wales/PHWC-16EAB6/2020\|EPI_ISL_585816\|2020 | ···················· | ···················· | ···················· | ···················· | ···················· | ···················· | |

| | | | | | | | | |
|---|---|---|---|---|---|---|---|---|
| M | 7960 | hCoV-19/Wales/PHWC-16EAC5/2020\|EPI_ISL_585817\|2020 | ···················· | ···················· | ···················· | ···················· | ···················· | ···················· |
| M | 7961 | hCoV-19/Wales/PHWC-16EAD4/2020\|EPI_ISL_585818\|2020 | ···················· | ···················· | ···················· | ···················· | ···················· | ···················· |
| M | 7962 | hCoV-19/Wales/PHWC-16EAE3/2020\|EPI_ISL_585819\|2020 | ···················· | ···················· | ···················· | ···················· | ···················· | ···················· |
| M | 7963 | hCoV-19/Wales/PHWC-16EAF2/2020\|EPI_ISL_585820\|2020 | ···················· | ···················· | ···················· | ···················· | ···················· | ···················· |
| M | 7964 | hCoV-19/Wales/PHWC-16EB0E/2020\|EPI_ISL_585821\|2020 | ···················· | ···················· | ···················· | ···················· | ···················· | ···················· |
| M | 7965 | hCoV-19/Wales/PHWC-16EB1D/2020\|EPI_ISL_585822\|2020 | ···················· | ···················· | ···················· | ···················· | ···················· | ···················· |
| M | 7966 | hCoV-19/Wales/PHWC-16EB2C/2020\|EPI_ISL_585823\|2020 | ···················· | ···················· | ···················· | ···················· | ···················· | ···················· |
| M | 7967 | hCoV-19/Wales/PHWC-16EB3B/2020\|EPI_ISL_585824\|2020 | ···················· | ···················· | ···················· | ···················· | ···················· | ···················· |
| M | 7968 | hCoV-19/Wales/PHWC-16EB59/2020\|EPI_ISL_585826\|2020 | ···················· | ···················· | ···················· | ···················· | ···················· | ···················· |
| M | 7969 | hCoV-19/Wales/PHWC-16EB68/2020\|EPI_ISL_585827\|2020 | ···················· | ···················· | ···················· | ···················· | ···················· | ···················· |
| M | 7970 | hCoV-19/Wales/PHWC-16EB77/2020\|EPI_ISL_585828\|2020 | ···················· | ···················· | ···················· | ···················· | ···················· | ···················· |
| M | 7971 | hCoV-19/Wales/PHWC-16EB86/2020\|EPI_ISL_585829\|2020 | ···················· | ···················· | ···················· | ···················· | ···················· | ···················· |
| M | 7972 | hCoV-19/Wales/PHWC-16EB95/2020\|EPI_ISL_585830\|2020 | ···················· | ···················· | ···················· | ···················· | ···················· | ···················· |
| M | 7973 | hCoV-19/Wales/PHWC-16EBA4/2020\|EPI_ISL_585831\|2020 | ···················· | ···················· | ···················· | ···················· | ···················· | ···················· |
| M | 7974 | hCoV-19/Wales/PHWC-16EBB3/2020\|EPI_ISL_585832\|2020 | ···················· | ···················· | ···················· | ···················· | ···················· | ···················· |
| M | 7975 | hCoV-19/Wales/PHWC-16EBC2/2020\|EPI_ISL_585833\|2020 | ···················· | ···················· | ···················· | ···················· | ···················· | ···················· |
| M | 7976 | hCoV-19/Wales/PHWC-16EBD1/2020\|EPI_ISL_585834\|2020 | ···················· | ···················· | ···················· | ···················· | ···················· | ···················· |
| M | 7977 | hCoV-19/Wales/PHWC-16EBE0/2020\|EPI_ISL_585835\|2020 | ···················· | ···················· | ···················· | ···················· | ···················· | ···················· |
| M | 7978 | hCoV-19/Wales/PHWC-16EBFF/2020\|EPI_ISL_585836\|2020 | ···················· | ···················· | ···················· | ···················· | ···················· | ···················· |
| M | 7979 | hCoV-19/Wales/PHWC-16EC0B/2020\|EPI_ISL_585837\|2020 | ···················· | ···················· | ···················· | ···················· | ···················· | ···················· |
| M | 7980 | hCoV-19/Wales/PHWC-16EC1A/2020\|EPI_ISL_585838\|2020 | ···················· | ···················· | ···················· | ···················· | ···················· | ···················· |
| M | 7981 | hCoV-19/Wales/PHWC-16EC29/2020\|EPI_ISL_585839\|2020 | ···················· | ···················· | ···················· | ···················· | ···················· | ···················· |
| M | 7982 | hCoV-19/Wales/PHWC-16EC47/2020\|EPI_ISL_585841\|2020 | ···················· | ···················· | ···················· | ···················· | ···················· | ···················· |
| M | 7983 | hCoV-19/Wales/PHWC-16EC56/2020\|EPI_ISL_585842\|2020 | ···················· | ···················· | ···················· | ···················· | ···················· | ···················· |
| M | 7984 | hCoV-19/Wales/PHWC-16EC65/2020\|EPI_ISL_585843\|2020 | ···················· | ···················· | ···················· | ···················· | ···················· | ···················· |
| M | 7985 | hCoV-19/Wales/PHWC-16EC74/2020\|EPI_ISL_585844\|2020 | ···················· | ···················· | ···················· | ···················· | ···················· | ···················· |
| M | 7986 | hCoV-19/Wales/PHWC-16EC92/2020\|EPI_ISL_585846\|2020 | ···················· | ···················· | ···················· | ···················· | ···················· | ···················· |
| M | 7987 | hCoV-19/Wales/PHWC-16ECA1/2020\|EPI_ISL_585847\|2020 | ···················· | ···················· | ···················· | ···················· | ···················· | ···················· |
| M | 7988 | hCoV-19/Wales/PHWC-16ECB0/2020\|EPI_ISL_585848\|2020 | ···················· | ···················· | ···················· | ···················· | ···················· | ···················· |
| M | 7989 | hCoV-19/Wales/PHWC-16ECCF/2020\|EPI_ISL_585849\|2020 | ···················· | ···················· | ···················· | ···················· | ···················· | ···················· |
| M | 7990 | hCoV-19/Wales/PHWC-16ECDE/2020\|EPI_ISL_585850\|2020 | ···················· | ···················· | ···················· | ···················· | ···················· | ···················· |
| M | 7991 | hCoV-19/England/ALDP-9FD6A1/2020\|EPI_ISL_588000\|20 | ···················· | ···················· | ···················· | ···················· | ···················· | ···················· |
| M | 7992 | hCoV-19/England/ALDP-9FDDBB/2020\|EPI_ISL_588001\|20 | ···················· | ···················· | ···················· | ···················· | ···················· | ···················· |
| M | 7993 | hCoV-19/England/ALDP-9FD1BF/2020\|EPI_ISL_588003\|20 | ···················· | ···················· | ···················· | ···················· | ···················· | ···················· |
| M | 7994 | hCoV-19/England/ALDP-9FCBD0/2020\|EPI_ISL_588004\|20 | ···················· | ···················· | ···················· | ···················· | ···················· | ···················· |
| M | 7995 | hCoV-19/England/ALDP-9FD45C/2020\|EPI_ISL_588005\|20 | ···················· | ···················· | ···················· | ···················· | ···················· | ···················· |
| M | 7996 | hCoV-19/England/ALDP-9FDCFA/2020\|EPI_ISL_588006\|20 | ···················· | ···················· | ···················· | ···················· | ···················· | ···················· |
| M | 7997 | hCoV-19/England/ALDP-9FD01C/2020\|EPI_ISL_588008\|20 | ···················· | ···················· | ···················· | ···················· | ···················· | ···················· |
| M | 7998 | hCoV-19/England/ALDP-9FD47A/2020\|EPI_ISL_588009\|20 | ···················· | ···················· | ···················· | ···················· | ···················· | ···················· |
| M | 7999 | hCoV-19/England/ALDP-9FC8BB/2020\|EPI_ISL_588010\|20 | ···················· | ···················· | ···················· | ···················· | ···················· | ···················· |
| M | 8000 | hCoV-19/England/ALDP-9FDAD2/2020\|EPI_ISL_588012\|20 | ···················· | ···················· | ···················· | ···················· | ···················· | ···················· |
| M | 8001 | hCoV-19/England/ALDP-9FDE30/2020\|EPI_ISL_588013\|20 | ···K················ | ···················· | ···················· | ···················· | ···················· | ···················· |
| M | 8002 | hCoV-19/England/ALDP-9FCB94/2020\|EPI_ISL_588029\|20 | ···················· | ···················· | ···················· | ···················· | ···················· | ···················· |
| M | 8003 | hCoV-19/England/ALDP-9FD3E6/2020\|EPI_ISL_588030\|20 | ···················· | ···················· | ···················· | ···················· | ···················· | ···················· |
| M | 8004 | hCoV-19/England/ALDP-9FCC19/2020\|EPI_ISL_588032\|20 | ···················· | ···················· | ···················· | ···················· | ···················· | ···················· |
| M | 8005 | hCoV-19/England/ALDP-9FD3D7/2020\|EPI_ISL_588033\|20 | ···················· | ···················· | ···················· | ···················· | ···················· | ···················· |
| M | 8006 | hCoV-19/England/ALDP-9FCDAD/2020\|EPI_ISL_588034\|20 | ···················· | ···················· | ···················· | ···················· | ···················· | ···················· |
| M | 8007 | hCoV-19/England/ALDP-9FCBEF/2020\|EPI_ISL_588036\|20 | ···················· | ···················· | ···················· | ···················· | ···················· | ···················· |
| M | 8008 | hCoV-19/England/ALDP-9FDAF0/2020\|EPI_ISL_588037\|20 | ···················· | ···················· | ···················· | ···················· | ···················· | ···················· |
| M | 8009 | hCoV-19/England/ALDP-9FC9C7/2020\|EPI_ISL_588038\|20 | ···················· | ·A·················· | ···················· | ···················· | ···················· | ··A················· |
| M | 8010 | hCoV-19/England/ALDP-9FD00D/2020\|EPI_ISL_588039\|20 | ···················· | ···················· | ···················· | ···················· | ···················· | ···················· |
| M | 8011 | hCoV-19/England/ALDP-9FDE03/2020\|EPI_ISL_588040\|20 | ···················· | ···················· | ···················· | ···················· | ···················· | ···················· |
| M | 8012 | hCoV-19/England/ALDP-9FDA0F/2020\|EPI_ISL_588041\|20 | ···················· | ···················· | ···················· | ···················· | ···················· | ···················· |
| M | 8013 | hCoV-19/England/ALDP-9FD780/2020\|EPI_ISL_588042\|20 | ···················· | ···················· | ···················· | ···················· | ······T············· | ···················· |
| M | 8014 | hCoV-19/England/ALDP-9FD3F5/2020\|EPI_ISL_588043\|20 | ···················· | ·A·················· | ···················· | ···················· | ···················· | ···················· |
| M | 8015 | hCoV-19/England/ALDP-9FCF4D/2020\|EPI_ISL_588044\|20 | ···················· | ···················· | ···················· | ···················· | ···················· | ···················· |
| M | 8016 | hCoV-19/England/ALDP-9FDEF4/2020\|EPI_ISL_588045\|20 | ···················· | ···················· | ···················· | ···················· | ···················· | ···················· |
| M | 8017 | hCoV-19/England/ALDP-9FDFE2/2020\|EPI_ISL_588046\|20 | ···················· | ···················· | ···················· | ···················· | ···················· | ···················· |
| M | 8018 | hCoV-19/England/ALDP-9FD331/2020\|EPI_ISL_588047\|20 | ···················· | ···················· | ···················· | ···················· | ···················· | ···················· |
| M | 8019 | hCoV-19/England/ALDP-9FC763/2020\|EPI_ISL_588048\|20 | ···················· | ···················· | ···················· | ···················· | ···················· | ···················· |
| M | 8020 | hCoV-19/England/ALDP-9FD683/2020\|EPI_ISL_588049\|20 | ···················· | ···················· | ···················· | ···················· | ···················· | ···················· |
| M | 8021 | hCoV-19/England/QEUH-9FA4F5/2020\|EPI_ISL_588060\|20 | ···················· | ···················· | ···················· | ···················· | ···················· | ···················· |
| M | 8022 | hCoV-19/England/QEUH-9FAE06/2020\|EPI_ISL_588062\|20 | ···················· | ···················· | ···················· | ···················· | ···················· | ···················· |
| M | 8023 | hCoV-19/England/QEUH-9FA774/2020\|EPI_ISL_588063\|20 | ···················· | ···················· | ···················· | ···················· | ···················· | ···················· |
| M | 8024 | hCoV-19/England/QEUH-9FA4B9/2020\|EPI_ISL_588064\|20 | ···················· | ···················· | ···················· | ···················· | ···················· | ···················· |
| M | 8025 | hCoV-19/England/QEUH-9FA880/2020\|EPI_ISL_588065\|20 | ···················· | ···················· | ···················· | ···················· | ···················· | ···················· |
| M | 8026 | hCoV-19/England/QEUH-9FA4C8/2020\|EPI_ISL_588066\|20 | ···················· | ···················· | ···················· | ···················· | ···················· | ···················· |
| M | 8027 | hCoV-19/England/QEUH-9FA747/2020\|EPI_ISL_588068\|20 | ···················· | ···················· | ···················· | ···················· | ···················· | ···················· |
| M | 8028 | hCoV-19/England/QEUH-9FAC75/2020\|EPI_ISL_588090\|20 | ···················· | ···················· | ···················· | ···················· | ···················· | ···················· |
| M | 8029 | hCoV-19/England/QEUH-9FB4D6/2020\|EPI_ISL_588091\|20 | ···················· | ···················· | ···················· | ···················· | ···················· | ···················· |
| M | 8030 | hCoV-19/England/QEUH-9FAA4E/2020\|EPI_ISL_588092\|20 | ···················· | ···················· | ···················· | ···················· | ···················· | ···················· |
| M | 8031 | hCoV-19/England/QEUH-9FA47D/2020\|EPI_ISL_588093\|20 | ···················· | ···················· | ···················· | ···················· | ···················· | ···················· |
| M | 8032 | hCoV-19/England/QEUH-9FADBE/2020\|EPI_ISL_588094\|20 | ···················· | ···················· | ···················· | ···················· | ···················· | ···················· |
| M | 8033 | hCoV-19/England/QEUH-9FA56B/2020\|EPI_ISL_588095\|20 | ···················· | ···················· | ···················· | ···················· | ···················· | ···················· |
| M | 8034 | hCoV-19/England/QEUH-9FAA7B/2020\|EPI_ISL_588096\|20 | ···················· | ···················· | ···················· | ···················· | ···················· | ···················· |
| M | 8035 | hCoV-19/England/QEUH-9FA792/2020\|EPI_ISL_588097\|20 | ···················· | ···················· | ···················· | ···················· | ···················· | ···················· |
| M | 8036 | hCoV-19/England/QEUH-9FB2DC/2020\|EPI_ISL_588098\|20 | ···················· | ···················· | ···················· | ···················· | ···················· | ···················· |
| M | 8037 | hCoV-19/England/QEUH-9FB00F/2020\|EPI_ISL_588099\|20 | ···················· | ···················· | ···················· | ···················· | ···················· | ···················· |
| M | 8038 | hCoV-19/England/ALDP-9E5BF8/2020\|EPI_ISL_589300\|20 | ···················· | ···················· | ···················· | ···················· | ···················· | ···················· |
| M | 8039 | hCoV-19/England/ALDP-9ECE25/2020\|EPI_ISL_589301\|20 | ···················· | ···················· | ···················· | ···················· | ···················· | ···················· |

| | | | | | | | |
|---|---|---|---|---|---|---|---|
| M | 8040 hCoV-19/England/MILK-9ECC58/2020\|EPI_ISL_589304\|20 | .................... | .................... | .................... | .................... | .................... | .................... |
| M | 8041 hCoV-19/England/ALDP-9EDECA/2020\|EPI_ISL_589305\|20 | .................... | .................... | .................... | .................... | .................... | .................... |
| M | 8042 hCoV-19/England/ALDP-9EE3CA/2020\|EPI_ISL_589306\|20 | .................... | .................... | .................... | .................... | .................... | .................... |
| M | 8043 hCoV-19/England/ALDP-9ED5A7/2020\|EPI_ISL_589307\|20 | .................... | .................... | .................... | .................... | .................... | .................... |
| M | 8044 hCoV-19/England/ALDP-9EDC84/2020\|EPI_ISL_589308\|20 | .................... | .................... | .................... | .................... | .................... | .................... |
| M | 8045 hCoV-19/England/ALDP-9EDF30/2020\|EPI_ISL_589309\|20 | .................... | .................... | .................... | .................... | .................... | .................... |
| M | 8046 hCoV-19/England/ALDP-9EE069/2020\|EPI_ISL_589310\|20 | .................... | .................... | .................... | .................... | .................... | .................... |
| M | 8047 hCoV-19/England/ALDP-9EDB87/2020\|EPI_ISL_589311\|20 | .................... | .................... | .................... | .................... | .................... | .................... |
| M | 8048 hCoV-19/England/ALDP-9ECF40/2020\|EPI_ISL_589312\|20 | .................... | .................... | .................... | .................... | .................... | .................... |
| M | 8049 hCoV-19/England/ALDP-9ECDCE/2020\|EPI_ISL_589313\|20 | .................... | .................... | .................... | .................... | .................... | .................... |
| M | 8050 hCoV-19/England/ALDP-9ED2BF/2020\|EPI_ISL_589314\|20 | .................... | .................... | .................... | .................... | .................... | .................... |
| M | 8051 hCoV-19/England/ALDP-9E5ACE/2020\|EPI_ISL_589315\|20 | .................... | .................... | .................... | .................... | .................... | .................... |
| M | 8052 hCoV-19/England/ALDP-9EE3AC/2020\|EPI_ISL_589316\|20 | .................... | .................... | .................... | .................... | .................... | .................... |
| M | 8053 hCoV-19/England/ALDP-9E5C40/2020\|EPI_ISL_589317\|20 | .................... | .A.................. | .................... | .................... | .................... | ...A................ |
| M | 8054 hCoV-19/England/ALDP-9E5AFB/2020\|EPI_ISL_589318\|20 | .................... | .................... | .................... | .................... | .................... | .................... |
| M | 8055 hCoV-19/England/ALDP-9ED6D1/2020\|EPI_ISL_589319\|20 | .................... | .................... | .................... | .................... | .................... | .................... |
| M | 8056 hCoV-19/England/ALDP-9EE02D/2020\|EPI_ISL_589320\|20 | .................... | .................... | .................... | .................... | .................... | .................... |
| M | 8057 hCoV-19/England/ALDP-9ECF8C/2020\|EPI_ISL_589321\|20 | .................... | .................... | .................... | .................... | .................... | .................... |
| M | 8058 hCoV-19/England/ALDP-9ED3DA/2020\|EPI_ISL_589322\|20 | .................... | .................... | .................... | .................... | .................... | .................... |
| M | 8059 hCoV-19/England/ALDP-9EDC93/2020\|EPI_ISL_589323\|20 | .................... | .................... | .................... | .................... | .................... | .................... |
| M | 8060 hCoV-19/England/ALDP-9E590D/2020\|EPI_ISL_589324\|20 | .................... | .................... | .................... | .................... | .................... | .................... |
| M | 8061 hCoV-19/England/ALDP-9ECFF5/2020\|EPI_ISL_589326\|20 | .................... | .................... | .................... | .................... | .................... | .................... |
| M | 8062 hCoV-19/England/ALDP-9ED06A/2020\|EPI_ISL_589327\|20 | .................... | .................... | .................... | .................... | .................... | .................... |
| M | 8063 hCoV-19/England/ALDP-9E58B5/2020\|EPI_ISL_589328\|20 | .................... | .................... | .................... | .................... | .................... | .................... |
| M | 8064 hCoV-19/England/ALDP-9E5C6E/2020\|EPI_ISL_589330\|20 | .................... | .................... | .................... | .................... | .................... | .................... |
| M | 8065 hCoV-19/England/ALDP-9ED219/2020\|EPI_ISL_589331\|20 | .................... | .................... | .................... | .................... | .................... | .................... |
| M | 8066 hCoV-19/England/ALDP-9ED783/2020\|EPI_ISL_589332\|20 | .................... | .................... | .................... | .................... | .................... | .................... |
| M | 8067 hCoV-19/England/ALDP-9ED3BC/2020\|EPI_ISL_589333\|20 | .................... | .................... | .................... | .................... | .................... | .................... |
| M | 8068 hCoV-19/England/MILK-9ECD55/2020\|EPI_ISL_589335\|20 | .................... | .................... | .................... | .................... | .................... | .................... |
| M | 8069 hCoV-19/England/ALDP-9ED7FC/2020\|EPI_ISL_589336\|20 | .................... | .................... | .................... | .................... | .................... | .................... |
| M | 8070 hCoV-19/England/ALDP-9E57A9/2020\|EPI_ISL_589337\|20 | .................... | .................... | .................... | .................... | .................... | .................... |
| M | 8071 hCoV-19/England/ALDP-9EE324/2020\|EPI_ISL_589338\|20 | .................... | .................... | .................... | .................... | .................... | .................... |
| M | 8072 hCoV-19/England/ALDP-9EE2CD/2020\|EPI_ISL_589339\|20 | .................... | .................... | .................... | .................... | .................... | .................... |
| M | 8073 hCoV-19/England/ALDP-9E5721/2020\|EPI_ISL_589341\|20 | .................... | .................... | .................... | .................... | .................... | .................... |
| M | 8074 hCoV-19/England/ALDP-9EDF03/2020\|EPI_ISL_589343\|20 | .................... | .................... | .................... | .................... | .................... | .................... |
| M | 8075 hCoV-19/England/ALDP-9ECF6E/2020\|EPI_ISL_589360\|20 | .................... | .................... | .................... | .................... | .................... | .................... |
| M | 8076 hCoV-19/England/ALDP-9EE096/2020\|EPI_ISL_589361\|20 | .................... | .................... | .................... | .................... | .................... | .................... |
| M | 8077 hCoV-19/England/ALDP-9ED39E/2020\|EPI_ISL_589362\|20 | .................... | .A.................. | .................... | .................... | .................... | .................... |
| M | 8078 hCoV-19/England/ALDP-9ED695/2020\|EPI_ISL_589363\|20 | .................... | .................... | .................... | .................... | .................... | .................... |
| M | 8079 hCoV-19/England/ALDP-9ED158/2020\|EPI_ISL_589364\|20 | .................... | .................... | .................... | .................... | .................... | .................... |
| M | 8080 hCoV-19/England/ALDP-9ED237/2020\|EPI_ISL_589365\|20 | .................... | .................... | .................... | .................... | .................... | .................... |
| M | 8081 hCoV-19/England/ALDP-9ED255/2020\|EPI_ISL_589366\|20 | .................... | .................... | .................... | .................... | .................... | .................... |
| M | 8082 hCoV-19/England/ALDP-9E5703/2020\|EPI_ISL_589367\|20 | .................... | .................... | .................... | .................... | .................... | .................... |
| M | 8083 hCoV-19/England/ALDP-9EDDDC/2020\|EPI_ISL_589368\|20 | .................... | .................... | .................... | .................... | .................... | .................... |
| M | 8084 hCoV-19/England/ALDP-9ED598/2020\|EPI_ISL_589369\|20 | .................... | .................... | .................... | .................... | .................... | .................... |
| M | 8085 hCoV-19/England/ALDP-9E586A/2020\|EPI_ISL_589370\|20 | .................... | .................... | .................... | .................... | .................... | .................... |
| M | 8086 hCoV-19/England/ALDP-9ED2DD/2020\|EPI_ISL_589371\|20 | .................... | .................... | .................... | .................... | .................... | .................... |
| M | 8087 hCoV-19/England/ALDP-9E59D0/2020\|EPI_ISL_589372\|20 | .................... | .................... | .................... | .................... | .................... | .................... |
| M | 8088 hCoV-19/England/ALDP-9E578B/2020\|EPI_ISL_589373\|20 | .................... | .................... | .................... | .................... | .................... | .................... |
| M | 8089 hCoV-19/England/ALDP-9EE00F/2020\|EPI_ISL_589374\|20 | .................... | .................... | .................... | .................... | .................... | .................... |
| M | 8090 hCoV-19/England/ALDP-9ED7ED/2020\|EPI_ISL_589375\|20 | .................... | .................... | .................... | .................... | .................... | .................... |
| M | 8091 hCoV-19/England/MILK-9ECC3A/2020\|EPI_ISL_589377\|20 | .................... | .................... | .................... | .................... | .................... | .................... |
| M | 8092 hCoV-19/England/ALDP-9E57F4/2020\|EPI_ISL_589378\|20 | .................... | .................... | .................... | .................... | .................... | .................... |
| M | 8093 hCoV-19/England/ALDP-9EE0B4/2020\|EPI_ISL_589379\|20 | .................... | .................... | .................... | .................... | .................... | .................... |
| M | 8094 hCoV-19/England/ALDP-9EE342/2020\|EPI_ISL_589391\|20 | .................... | .................... | .................... | .................... | .................... | .................... |
| M | 8095 hCoV-19/England/ALDP-9ED4C8/2020\|EPI_ISL_589392\|20 | .................... | .................... | .................... | .................... | .................... | .................... |
| M | 8096 hCoV-19/England/ALDP-9EE139/2020\|EPI_ISL_589393\|20 | .................... | .................... | .................... | .................... | .................... | .................... |
| M | 8097 hCoV-19/England/ALDP-9E5985/2020\|EPI_ISL_589394\|20 | .................... | .................... | .................... | .................... | .................... | .................... |
| M | 8098 hCoV-19/England/ALDP-9ECDEC/2020\|EPI_ISL_589395\|20 | .................... | .................... | .................... | .................... | .................... | .................... |
| M | 8099 hCoV-19/England/ALDP-9ED0B5/2020\|EPI_ISL_589396\|20 | .................... | .................... | .................... | .................... | .................... | .................... |
| M | 8100 hCoV-19/England/ALDP-9EE351/2020\|EPI_ISL_589397\|20 | .................... | .................... | .................... | .................... | .................... | .................... |
| M | 8101 hCoV-19/England/ALDP-9E5958/2020\|EPI_ISL_589398\|20 | .................... | .................... | .................... | .................... | .................... | .................... |
| M | 8102 hCoV-19/England/ALDP-9B5960/2020\|EPI_ISL_590300\|20 | .................... | .................... | .................... | .................... | .................... | .................... |
| M | 8103 hCoV-19/England/QEUH-9B4C86/2020\|EPI_ISL_590301\|20 | .................... | .................... | .................... | .................... | .................... | .................... |
| M | 8104 hCoV-19/England/QEUH-9B46A6/2020\|EPI_ISL_590302\|20 | .................... | .................... | .................... | .................... | ....T............... | .................... |
| M | 8105 hCoV-19/England/QEUH-9B5414/2020\|EPI_ISL_590303\|20 | .................... | .................... | .................... | .................... | .................... | .................... |
| M | 8106 hCoV-19/England/QEUH-9B565A/2020\|EPI_ISL_590304\|20 | .................... | .................... | .................... | .................... | .................... | .................... |
| M | 8107 hCoV-19/England/QEUH-9B56B4/2020\|EPI_ISL_590305\|20 | .................... | .................... | .................... | .................... | .................... | .................... |
| M | 8108 hCoV-19/England/ALDP-9B57B1/2020\|EPI_ISL_590306\|20 | .................... | .................... | .................... | .................... | .................... | .................... |
| M | 8109 hCoV-19/England/ALDP-9B5793/2020\|EPI_ISL_590309\|20 | .................... | .................... | .................... | .................... | .................... | .................... |
| M | 8110 hCoV-19/England/QEUH-9B4D1A/2020\|EPI_ISL_590310\|20 | .................... | .................... | .................... | .................... | .................... | .................... |
| M | 8111 hCoV-19/England/QEUH-9B4B98/2020\|EPI_ISL_590311\|20 | .................... | .................... | .................... | .................... | .................... | .................... |
| M | 8112 hCoV-19/England/QEUH-9B554E/2020\|EPI_ISL_590313\|20 | .................... | .................... | .................... | .................... | .................... | .................... |
| M | 8113 hCoV-19/England/QEUH-9B4F14/2020\|EPI_ISL_590315\|20 | .................... | .................... | .................... | .................... | .................... | .................... |
| M | 8114 hCoV-19/England/QEUH-9B4DCF/2020\|EPI_ISL_590316\|20 | .................... | .................... | .................... | .................... | .................... | .................... |
| M | 8115 hCoV-19/England/QEUH-9B5371/2020\|EPI_ISL_590317\|20 | .................... | .................... | .................... | .................... | .................... | .................... |
| M | 8116 hCoV-19/England/QEUH-9B4C4A/2020\|EPI_ISL_590318\|20 | .................... | .................... | .................... | .................... | .................... | .................... |
| M | 8117 hCoV-19/England/ALDP-9B5809/2020\|EPI_ISL_590319\|20 | .................... | .................... | .................... | .................... | .................... | .................... |
| M | 8118 hCoV-19/England/ALDP-9B5854/2020\|EPI_ISL_590321\|20 | .................... | .................... | .................... | .................... | .................... | .................... |
| M | 8119 hCoV-19/England/QEUH-9B4BA7/2020\|EPI_ISL_590322\|20 | .................... | .................... | .................... | .................... | ......T............. | .................... |

| | | | | | | | | |
|---|---|---|---|---|---|---|---|---|
| M | 8120 hCoV-19/England/QEUH-9B4A13/2020\|EPI_ISL_590323\|20 | ···················· | ···················· | ···················· | ···················· | ···················· | ···················· | |
| M | 8121 hCoV-19/England/QEUH-9B4EEA/2020\|EPI_ISL_590324\|20 | ···················· | ···················· | ···················· | ···················· | ···················· | ···················· | |
| M | 8122 hCoV-19/England/QEUH-9B4846/2020\|EPI_ISL_590340\|20 | ···················· | ···················· | ···················· | ···················· | ···················· | ···················· | |
| M | 8123 hCoV-19/England/QEUH-9B4E53/2020\|EPI_ISL_590341\|20 | ···················· | ·A·················· | ···················· | ···················· | ···················· | ··A················· | |
| M | 8124 hCoV-19/England/QEUH-9B4F9C/2020\|EPI_ISL_590342\|20 | ···················· | ···················· | ···················· | ···················· | ···T················ | ···················· | |
| M | 8125 hCoV-19/England/QEUH-9B4D74/2020\|EPI_ISL_590343\|20 | ···················· | ···················· | ···················· | ···················· | ···················· | ···················· | |
| M | 8126 hCoV-19/England/QEUH-9B4EAE/2020\|EPI_ISL_590344\|20 | ···················· | ·A·················· | ···················· | ···················· | ···················· | ··A················· | |
| M | 8127 hCoV-19/England/QEUH-9B5317/2020\|EPI_ISL_590345\|20 | ···················· | ···················· | ···················· | ···················· | ···················· | ···················· | |
| M | 8128 hCoV-19/England/ALDP-9B5818/2020\|EPI_ISL_590346\|20 | ···················· | ···················· | ···················· | ···················· | ···················· | ···················· | |
| M | 8129 hCoV-19/England/ALDP-9B57DF/2020\|EPI_ISL_590347\|20 | ···················· | ···················· | ···················· | ···················· | ···················· | ···················· | |
| M | 8130 hCoV-19/England/QEUH-9B51B3/2020\|EPI_ISL_590348\|20 | ···················· | ·A·················· | ···················· | ···················· | ···················· | ··A················· | |
| M | 8131 hCoV-19/England/QEUH-9B4855/2020\|EPI_ISL_590349\|20 | ···················· | ···················· | ···················· | ···················· | ·····T·············· | ···················· | |
| M | 8132 hCoV-19/England/QEUH-9B4C0E/2020\|EPI_ISL_590350\|20 | ···················· | ···················· | ···················· | ···················· | ···················· | ···················· | |
| M | 8133 hCoV-19/England/ALDP-9B599D/2020\|EPI_ISL_590351\|20 | ···················· | ···················· | ···················· | ···················· | ···················· | ···················· | |
| M | 8134 hCoV-19/England/ALDP-9B5933/2020\|EPI_ISL_590352\|20 | ···················· | ···················· | ···················· | ···················· | ···················· | ···················· | |
| M | 8135 hCoV-19/England/QEUH-9B553F/2020\|EPI_ISL_590353\|20 | ···················· | ···················· | ···················· | ···················· | ···················· | ···················· | |
| M | 8136 hCoV-19/England/QEUH-9B5599/2020\|EPI_ISL_590354\|20 | ···················· | ···················· | ···················· | ···················· | ···················· | ···················· | |
| M | 8137 hCoV-19/England/ALDP-9B5748/2020\|EPI_ISL_590355\|20 | ···················· | ···················· | ···················· | ···················· | ···················· | ···················· | |
| M | 8138 hCoV-19/England/QEUH-9B4916/2020\|EPI_ISL_590356\|20 | ···················· | ···················· | ···················· | ···················· | ···················· | ···················· | |
| M | 8139 hCoV-19/England/ALDP-9B5BC4/2020\|EPI_ISL_590357\|20 | ···················· | ···················· | ···················· | ···················· | ···················· | ···················· | |
| M | 8140 hCoV-19/England/QEUH-9B4FE7/2020\|EPI_ISL_590358\|20 | ···················· | ···················· | ···················· | ···················· | ···················· | ···················· | |
| M | 8141 hCoV-19/England/QEUH-9B50E3/2020\|EPI_ISL_590360\|20 | ···················· | ···················· | ···················· | ···················· | ···················· | ···················· | |
| M | 8142 hCoV-19/England/QEUH-9B55A8/2020\|EPI_ISL_590372\|20 | ···················· | ···················· | ···················· | ···················· | ···················· | ···················· | |
| M | 8143 hCoV-19/England/ALDP-9B5C49/2020\|EPI_ISL_590373\|20 | ···················· | ···················· | ···················· | ···················· | ···················· | ···················· | |
| M | 8144 hCoV-19/England/QEUH-9B4FF6/2020\|EPI_ISL_590374\|20 | ···················· | ···················· | ···················· | ···················· | ···················· | ···················· | |
| M | 8145 hCoV-19/England/QEUH-9B4C59/2020\|EPI_ISL_590375\|20 | ···················· | ···················· | ···················· | ···················· | ···················· | ···················· | |
| M | 8146 hCoV-19/England/QEUH-9B514A/2020\|EPI_ISL_590378\|20 | ···················· | ···················· | ···················· | ···················· | ···T················ | ···················· | |
| M | 8147 hCoV-19/England/ALDP-9B5CB2/2020\|EPI_ISL_590379\|20 | ···················· | ···················· | ···················· | ···················· | ···················· | ···················· | |
| M | 8149 hCoV-19/Australia/VIC14662/2020\|EPI_ISL_592941\|202 | ···················· | ···················· | ···················· | ···················· | ···················· | ···················· | |
| M | 8151 hCoV-19/Australia/VIC14664/2020\|EPI_ISL_592943\|202 | ···················· | ···················· | ···················· | ···················· | ···················· | ···················· | |
| M | 8152 hCoV-19/Australia/VIC14665/2020\|EPI_ISL_592944\|202 | ···················· | ···················· | ···················· | ···················· | ···················· | ···················· | |
| M | 8153 hCoV-19/Australia/VIC14667/2020\|EPI_ISL_592945\|202 | ···················· | ···················· | ···················· | ···················· | ···················· | ···················· | |
| M | 8154 hCoV-19/Australia/VIC14670/2020\|EPI_ISL_592946\|202 | ···················· | ···················· | ···················· | ···················· | ···················· | ···················· | |
| M | 8156 hCoV-19/Australia/VIC14673/2020\|EPI_ISL_592948\|202 | ···················· | ···················· | ···················· | ···················· | ···················· | ···················· | |
| M | 8157 hCoV-19/Australia/VIC14675/2020\|EPI_ISL_592949\|202 | ···················· | ···················· | ···················· | ···················· | ···················· | ···················· | |
| M | 8158 hCoV-19/Australia/VIC14677/2020\|EPI_ISL_592950\|202 | ···················· | ···················· | ···················· | ···················· | ···················· | ···················· | |
| M | 8159 hCoV-19/Australia/VIC14681/2020\|EPI_ISL_592951\|202 | ···················· | ···················· | ···················· | ···················· | ···················· | ···················· | |
| M | 8160 hCoV-19/Australia/VIC14685/2020\|EPI_ISL_592952\|202 | ···················· | ···················· | ···················· | ···················· | ···················· | ···················· | |
| M | 8161 hCoV-19/Australia/VIC14686/2020\|EPI_ISL_592953\|202 | ···················· | ···················· | ···················· | ···················· | ···················· | ···················· | |
| M | 8162 hCoV-19/Australia/VIC14689/2020\|EPI_ISL_592954\|202 | ···················· | ···················· | ···················· | ···················· | ···················· | NNN················· | 1 |
| M | 8163 hCoV-19/Australia/VIC14690/2020\|EPI_ISL_592955\|202 | ···················· | ···················· | ···················· | ···················· | ···················· | ···················· | |
| M | 8164 hCoV-19/Australia/VIC14692/2020\|EPI_ISL_592956\|202 | ···················· | ···················· | ···················· | ···················· | ···················· | ···················· | |
| M | 8165 hCoV-19/Australia/VIC14698/2020\|EPI_ISL_592957\|202 | ···················· | ···················· | ···················· | ···················· | ···················· | ···················· | |
| M | 8166 hCoV-19/Australia/VIC14700/2020\|EPI_ISL_592958\|202 | ···················· | ···················· | ···K················ | ···················· | ···················· | ···················· | |
| M | 8168 hCoV-19/Australia/VIC14704/2020\|EPI_ISL_592960\|202 | ···················· | ···················· | ···················· | ···················· | ···················· | ···················· | |
| M | 8169 hCoV-19/Australia/VIC14706/2020\|EPI_ISL_592961\|202 | ···················· | ···················· | ···················· | ···················· | ···················· | ···················· | |
| M | 8170 hCoV-19/Australia/VIC14708/2020\|EPI_ISL_592962\|202 | ···················· | ···················· | ···················· | ···················· | ···················· | ···················· | |
| M | 8173 hCoV-19/Australia/VIC14713/2020\|EPI_ISL_592965\|202 | ···················· | ···················· | ···················· | ···················· | ···················· | ···················· | |
| M | 8174 hCoV-19/Australia/VIC14715/2020\|EPI_ISL_592966\|202 | ···················· | ···················· | ···················· | ···················· | ···················· | ···················· | |
| M | 8176 hCoV-19/Australia/VIC14719/2020\|EPI_ISL_592968\|202 | ···················· | ···················· | ···················· | ···················· | ···················· | ···················· | |
| M | 8177 hCoV-19/Australia/VIC14722/2020\|EPI_ISL_592969\|202 | ···················· | ···················· | ···················· | ···················· | ···················· | ···················· | |
| M | 8178 hCoV-19/Australia/VIC14725/2020\|EPI_ISL_592970\|202 | ···················· | ···················· | ···················· | ···················· | ···················· | ···················· | |
| M | 8179 hCoV-19/Australia/VIC14727/2020\|EPI_ISL_592971\|202 | ···················· | ···················· | ···K················ | ···················· | ···················· | ···················· | |
| M | 8180 hCoV-19/Australia/VIC14728/2020\|EPI_ISL_592972\|202 | ···················· | ···················· | ···················· | ···················· | ···················· | ···················· | |
| M | 8182 hCoV-19/Australia/VIC14731/2020\|EPI_ISL_592974\|202 | ···················· | ···················· | ···················· | ···················· | ···················· | ···················· | |
| M | 8183 hCoV-19/Australia/VIC14734/2020\|EPI_ISL_592975\|202 | ···················· | ···················· | ···K················ | ···················· | ···················· | ···················· | |
| M | 8184 hCoV-19/Australia/VIC14735/2020\|EPI_ISL_592976\|202 | ···················· | ···················· | ···················· | ···················· | ···················· | ···················· | |
| M | 8185 hCoV-19/Australia/VIC14737/2020\|EPI_ISL_592977\|202 | ···················· | ···················· | ···················· | ···················· | ···················· | ···················· | |
| M | 8186 hCoV-19/Australia/VIC14738/2020\|EPI_ISL_592978\|202 | ···················· | ···················· | ···················· | ···················· | ···················· | ···················· | |
| M | 8187 hCoV-19/Australia/VIC14740/2020\|EPI_ISL_592979\|202 | ···················· | ···················· | ···················· | ···················· | ···················· | ···················· | |
| M | 8188 hCoV-19/Australia/VIC14744/2020\|EPI_ISL_592981\|202 | ···················· | ···················· | ···················· | ···················· | ···················· | ···················· | |
| M | 8189 hCoV-19/Australia/VIC14747/2020\|EPI_ISL_592983\|202 | ···················· | ···················· | ···················· | ···················· | ···················· | ···················· | |
| M | 8190 hCoV-19/Australia/VIC14748/2020\|EPI_ISL_592984\|202 | ···················· | ···················· | ···················· | ···················· | ···················· | ···················· | |
| M | 8191 hCoV-19/Australia/VIC14749/2020\|EPI_ISL_592985\|202 | ···················· | ···················· | ···················· | ···················· | ···················· | ···················· | |
| M | 8192 hCoV-19/Australia/VIC14756/2020\|EPI_ISL_592986\|202 | ···················· | ···················· | ···················· | ···················· | ···················· | ···················· | |
| M | 8193 hCoV-19/England/CWAR-24FDD9C/2020\|EPI_ISL_584521\|2 | ···················· | ···················· | ···················· | ···················· | ···················· | ···················· | |
| M | 8194 hCoV-19/England/CWAR-24FDDAB/2020\|EPI_ISL_584522\|2 | ···················· | ···················· | ···················· | ···················· | ···················· | ···················· | |
| M | 8195 hCoV-19/England/CWAR-24FDDBA/2020\|EPI_ISL_584523\|2 | ···················· | ···················· | ···················· | ···················· | ···················· | ···················· | |
| M | 8196 hCoV-19/England/CWAR-24FDDC9/2020\|EPI_ISL_584524\|2 | ···················· | ···················· | ···················· | ···················· | ···················· | ···················· | |
| M | 8197 hCoV-19/England/CWAR-24FDDD8/2020\|EPI_ISL_584525\|2 | ···················· | ···················· | ···················· | ···················· | ···················· | ···················· | |
| M | 8198 hCoV-19/England/CWAR-24FDDE7/2020\|EPI_ISL_584526\|2 | ···················· | ···················· | ···················· | ···················· | ···················· | ···················· | |
| M | 8199 hCoV-19/England/CWAR-24FDE02/2020\|EPI_ISL_584528\|2 | ···················· | ···A················ | ···················· | ···················· | ···················· | ···················· | |
| M | 8200 hCoV-19/England/CWAR-24FDE11/2020\|EPI_ISL_584529\|2 | ···················· | ···················· | ···················· | ···················· | ···················· | ···················· | |
| M | 8201 hCoV-19/England/CWAR-24FDE20/2020\|EPI_ISL_584530\|2 | ···················· | ···················· | ···················· | ···················· | ···················· | ···················· | |
| M | 8202 hCoV-19/England/CWAR-24FDE4E/2020\|EPI_ISL_584532\|2 | ···················· | ···················· | ···················· | ···················· | ···················· | ···················· | |
| M | 8203 hCoV-19/England/CWAR-24FDE7B/2020\|EPI_ISL_584535\|2 | ···················· | ···················· | ···················· | ···················· | ···················· | ···················· | |
| M | 8204 hCoV-19/England/CWAR-24FDE8A/2020\|EPI_ISL_584536\|2 | ···················· | ···················· | ···················· | ···················· | ···················· | ···················· | |
| M | 8205 hCoV-19/England/CWAR-24FDE99/2020\|EPI_ISL_584537\|2 | ···················· | ···················· | ···················· | ···················· | ···················· | ···················· | |
| M | 8206 hCoV-19/England/CWAR-24FDEA8/2020\|EPI_ISL_584538\|2 | ···················· | ···A················ | ···················· | ···················· | ···················· | ···················· | |
| M | 8207 hCoV-19/England/CWAR-24FDEC6/2020\|EPI_ISL_584540\|2 | ···················· | ···················· | ···················· | ···················· | ···················· | ···················· | |

| | | | | | | | | | |
|---|---|---|---|---|---|---|---|---|---|
| M | 8208 | hCoV-19/England/CWAR-24FDEE4/2020\|EPI_ISL_584542\|2 | ···················· | ···················· | ····················· | ···················· | ···················· | ···················· | |
| M | 8209 | hCoV-19/England/CWAR-24FDEF3/2020\|EPI_ISL_584543\|2 | ···················· | ···A················ | ····················· | ···················· | ···················· | ···················· | |
| M | 8210 | hCoV-19/England/CWAR-24FDF0F/2020\|EPI_ISL_584544\|2 | ···················· | ···················· | ····················· | ···················· | ···················· | ···················· | |
| M | 8211 | hCoV-19/England/CWAR-24FDF1E/2020\|EPI_ISL_584545\|2 | ···················· | ···················· | ····················· | ···················· | ···················· | ···················· | |
| M | 8212 | hCoV-19/England/CWAR-24FDF2D/2020\|EPI_ISL_584546\|2 | ···················· | ···················· | ····················· | ···················· | ···················· | ···················· | |
| M | 8213 | hCoV-19/England/CWAR-24FDF3C/2020\|EPI_ISL_584547\|2 | ···················· | ···················· | ····················· | ···················· | ···················· | ···················· | |
| M | 8214 | hCoV-19/England/CWAR-24FDF4B/2020\|EPI_ISL_584548\|2 | ···················· | ···················· | ····················· | ···················· | ···················· | ···················· | |
| M | 8215 | hCoV-19/England/CWAR-24FDF5A/2020\|EPI_ISL_584549\|2 | ···················· | ···················· | ····················· | ···················· | ···················· | ···················· | |
| M | 8216 | hCoV-19/England/CWAR-24FDF69/2020\|EPI_ISL_584550\|2 | ···················· | ···A················ | ····················· | ···················· | ···················· | ···················· | |
| M | 8217 | hCoV-19/England/CWAR-24FDF87/2020\|EPI_ISL_584552\|2 | ···················· | ···················· | ····················· | ···················· | ···················· | ···················· | |
| M | 8218 | hCoV-19/England/CWAR-24FDFA5/2020\|EPI_ISL_584554\|2 | ···················· | ···················· | ····················· | ···················· | ···················· | ···················· | |
| M | 8219 | hCoV-19/England/CWAR-24FDFB4/2020\|EPI_ISL_584555\|2 | ···················· | ···················· | ····················· | ···················· | ···················· | ···················· | |
| M | 8220 | hCoV-19/England/CWAR-24FDFC3/2020\|EPI_ISL_584556\|2 | ···················· | ···················· | ····················· | ···················· | ···················· | ···················· | |
| M | 8221 | hCoV-19/England/CWAR-24FE00B/2020\|EPI_ISL_584560\|2 | ···················· | ···················· | ····················· | ···················· | ···················· | ···················· | |
| M | 8222 | hCoV-19/England/CWAR-24FE029/2020\|EPI_ISL_584562\|2 | ···················· | ···················· | ····················· | ···················· | ···················· | ···················· | |
| M | 8223 | hCoV-19/England/CWAR-24FE038/2020\|EPI_ISL_584563\|2 | ···················· | ···················· | ····················· | ···················· | ···················· | ···················· | |
| M | 8224 | hCoV-19/England/CWAR-24FE047/2020\|EPI_ISL_584564\|2 | ···················· | ···A················ | ····················· | ···················· | ···················· | ···················· | |
| M | 8225 | hCoV-19/England/CWAR-24FE065/2020\|EPI_ISL_584566\|2 | ···················· | ···················· | ····················· | ···················· | ···················· | ···················· | |
| M | 8226 | hCoV-19/England/CWAR-24FE074/2020\|EPI_ISL_584567\|2 | ···················· | ···················· | ····················· | ···················· | ···················· | ···················· | |
| M | 8227 | hCoV-19/England/CWAR-24FE083/2020\|EPI_ISL_584568\|2 | ···················· | ···················· | ····················· | ···················· | ···················· | ···················· | |
| M | 8228 | hCoV-19/England/CWAR-24FE0CF/2020\|EPI_ISL_584571\|2 | ···················· | ···················· | ····················· | ···················· | ···················· | ···················· | |
| M | 8229 | hCoV-19/England/CWAR-24FE0FC/2020\|EPI_ISL_584573\|2 | ···················· | ···················· | ····················· | ···················· | ···················· | ···················· | |
| M | 8230 | hCoV-19/England/CWAR-24FE108/2020\|EPI_ISL_584574\|2 | ···················· | ···················· | ····················· | ···················· | ···················· | ···················· | |
| M | 8232 | hCoV-19/England/CWAR-24FE171/2020\|EPI_ISL_584579\|2 | ···················· | ···················· | ····················· | ···················· | ···················· | ···················· | |
| M | 8233 | hCoV-19/England/CWAR-24FE19F/2020\|EPI_ISL_584581\|2 | ···················· | ···················· | ····················· | ···················· | ···················· | ···················· | |
| M | 8234 | hCoV-19/England/CWAR-24FE1AE/2020\|EPI_ISL_584582\|2 | ···················· | ···················· | ····················· | ···················· | ···················· | ···················· | |
| M | 8235 | hCoV-19/England/CWAR-24FE1CC/2020\|EPI_ISL_584584\|2 | ···················· | ···················· | ····················· | ···················· | ···················· | ···················· | |
| M | 8236 | hCoV-19/Wales/PHWC-16ECED/2020\|EPI_ISL_585851\|2020 | ···················· | ···················· | ····················· | ···················· | ···················· | ···················· | |
| M | 8237 | hCoV-19/Wales/PHWC-16ECFC/2020\|EPI_ISL_585852\|2020 | ···················· | ···················· | ····················· | ···················· | ···················· | ···················· | |
| M | 8238 | hCoV-19/Wales/PHWC-16ED26/2020\|EPI_ISL_585855\|2020 | ···················· | ···················· | ····················· | ···················· | ···················· | ···················· | |
| M | 8239 | hCoV-19/Wales/PHWC-16ED35/2020\|EPI_ISL_585856\|2020 | ···················· | ···················· | ····················· | ···················· | ···················· | ···················· | |
| M | 8240 | hCoV-19/Wales/PHWC-16ED44/2020\|EPI_ISL_585857\|2020 | ···················· | ···················· | ····················· | ···················· | ···················· | ···················· | |
| M | 8242 | hCoV-19/Wales/PHWC-16ED71/2020\|EPI_ISL_585859\|2020 | ···················· | ···················· | ····················· | ···················· | ···················· | ···················· | |
| M | 8243 | hCoV-19/Wales/PHWC-16ED80/2020\|EPI_ISL_585860\|2020 | ···················· | ···················· | ····················· | ···················· | ···················· | ···················· | |
| M | 8244 | hCoV-19/Wales/PHWC-16ED9F/2020\|EPI_ISL_585861\|2020 | ···················· | ···················· | ····················· | ···················· | ···················· | ···················· | |
| M | 8245 | hCoV-19/Wales/PHWC-16EDAE/2020\|EPI_ISL_585862\|2020 | ···················· | ···················· | ····················· | ···················· | ···················· | ···················· | |
| M | 8246 | hCoV-19/Wales/PHWC-16EDBD/2020\|EPI_ISL_585863\|2020 | ···················· | ···················· | ····················· | ···················· | ···················· | ···················· | |
| M | 8247 | hCoV-19/Wales/PHWC-16EDCC/2020\|EPI_ISL_585864\|2020 | ···················· | ···················· | ····················· | ···················· | ···················· | ···················· | |
| M | 8248 | hCoV-19/Wales/PHWC-16EDDB/2020\|EPI_ISL_585865\|2020 | ···················· | ···················· | ····················· | ···················· | ···················· | ···················· | |
| M | 8249 | hCoV-19/Wales/PHWC-16EDEA/2020\|EPI_ISL_585866\|2020 | ···················· | ···················· | ····················· | ···················· | ···················· | ···················· | |
| M | 8250 | hCoV-19/Wales/PHWC-16EDF9/2020\|EPI_ISL_585867\|2020 | ···················· | ···················· | ····················· | ···················· | ···················· | ···················· | |
| M | 8251 | hCoV-19/Wales/PHWC-16EE14/2020\|EPI_ISL_585869\|2020 | ···················· | ···················· | ····················· | ···················· | ···················· | ···················· | |
| M | 8252 | hCoV-19/Wales/PHWC-16EE23/2020\|EPI_ISL_585870\|2020 | ···················· | ···················· | ····················· | ···················· | ···················· | ···················· | |
| M | 8253 | hCoV-19/Wales/PHWC-16EE32/2020\|EPI_ISL_585871\|2020 | ···················· | ···················· | ····················· | ···················· | ···················· | ···················· | |
| M | 8254 | hCoV-19/Wales/PHWC-16EE50/2020\|EPI_ISL_585873\|2020 | ···················· | ···················· | ····················· | ···················· | ···················· | ···················· | |
| M | 8255 | hCoV-19/Wales/PHWC-16EE7E/2020\|EPI_ISL_585875\|2020 | ···················· | ···················· | ····················· | ···················· | ···················· | ···················· | |
| M | 8256 | hCoV-19/Wales/PHWC-16EE8D/2020\|EPI_ISL_585876\|2020 | ···················· | ···················· | ····················· | ···················· | ···················· | ···················· | |
| M | 8257 | hCoV-19/Wales/PHWC-16EEAB/2020\|EPI_ISL_585878\|2020 | ···················· | ···················· | ····················· | ···················· | ···················· | ···················· | |
| M | 8258 | hCoV-19/Wales/PHWC-16EEBA/2020\|EPI_ISL_585879\|2020 | ···················· | ···················· | ····················· | ···················· | ···················· | ···················· | |
| M | 8259 | hCoV-19/Wales/PHWC-16EEC9/2020\|EPI_ISL_585880\|2020 | ···················· | ···················· | ····················· | ···················· | ···················· | ···················· | |
| M | 8260 | hCoV-19/Wales/PHWC-16EED8/2020\|EPI_ISL_585881\|2020 | ···················· | ···················· | ····················· | ···················· | ···················· | ···················· | |
| M | 8261 | hCoV-19/Wales/PHWC-16EEE7/2020\|EPI_ISL_585882\|2020 | ···················· | ···················· | ····················· | ···················· | ···················· | ···················· | |
| M | 8262 | hCoV-19/Wales/PHWC-16EEF6/2020\|EPI_ISL_585883\|2020 | ···················· | ···················· | ····················· | ···················· | ···················· | ···················· | |
| M | 8263 | hCoV-19/Wales/PHWC-16EF02/2020\|EPI_ISL_585884\|2020 | ···················· | ···················· | ····················· | ···················· | ···················· | ···················· | |
| M | 8265 | hCoV-19/Wales/PHWC-16EF20/2020\|EPI_ISL_585886\|2020 | ···················· | ···················· | ····················· | ···················· | ···················· | ···················· | |
| M | 8266 | hCoV-19/Wales/PHWC-16EF3F/2020\|EPI_ISL_585887\|2020 | ···················· | ···················· | ····················· | ···················· | ···················· | ···················· | |
| M | 8267 | hCoV-19/Wales/PHWC-16EF7B/2020\|EPI_ISL_585891\|2020 | ···················· | ···················· | ····················· | ···················· | ···················· | ···················· | |
| M | 8268 | hCoV-19/Wales/PHWC-16EF8A/2020\|EPI_ISL_585892\|2020 | ···················· | ···················· | ····················· | ···················· | ···················· | ···················· | |
| M | 8269 | hCoV-19/Wales/PHWC-16EF99/2020\|EPI_ISL_585893\|2020 | ···················· | ···················· | ····················· | ···················· | ···················· | ···················· | |
| M | 8270 | hCoV-19/Wales/PHWC-16EFA8/2020\|EPI_ISL_585894\|2020 | ···················· | ···················· | ····················· | ···················· | ···················· | ···················· | |
| M | 8271 | hCoV-19/Wales/PHWC-16EFB7/2020\|EPI_ISL_585895\|2020 | ···················· | ···················· | ····················· | ···················· | ···················· | ···················· | |
| M | 8272 | hCoV-19/Wales/PHWC-16EFC6/2020\|EPI_ISL_585896\|2020 | ···················· | ···················· | ····················· | ···················· | ···················· | ···················· | |
| M | 8273 | hCoV-19/Wales/PHWC-16EFD5/2020\|EPI_ISL_585897\|2020 | ···················· | ···················· | ····················· | ···················· | ···················· | ···················· | |
| M | 8274 | hCoV-19/Wales/PHWC-16EFF3/2020\|EPI_ISL_585898\|2020 | ···················· | ···················· | ····················· | ···················· | ···················· | ···················· | |
| M | 8275 | hCoV-19/Wales/PHWC-16F00E/2020\|EPI_ISL_585899\|2020 | ···················· | ···················· | ····················· | ···················· | ···················· | ···················· | |
| M | 8276 | hCoV-19/England/ALDP-9FDDD9/2020\|EPI_ISL_588014\|20 | ···················· | ···················· | ····················· | ···················· | ···················· | ···················· | |
| M | 8277 | hCoV-19/England/ALDP-9FDAB4/2020\|EPI_ISL_588015\|20 | ···················· | ···················· | ····················· | ···················· | ···················· | ···················· | |
| M | 8278 | hCoV-19/England/ALDP-9FCCA0/2020\|EPI_ISL_588016\|20 | ···················· | ···················· | ····················· | ···················· | ···················· | ···················· | |
| M | 8279 | hCoV-19/England/ALDP-9FDEA9/2020\|EPI_ISL_588018\|20 | ···················· | ···················· | ····················· | ···················· | ······T············· | ···················· | |
| M | 8280 | hCoV-19/England/ALDP-9FCFE3/2020\|EPI_ISL_588020\|20 | ···················· | ···················· | ····················· | ···················· | ···················· | ···················· | |
| M | 8281 | hCoV-19/England/ALDP-9FCC37/2020\|EPI_ISL_588021\|20 | ···················· | ···················· | ····················· | ···················· | ···················· | ···················· | |
| M | 8282 | hCoV-19/England/ALDP-9FC9F4/2020\|EPI_ISL_588022\|20 | ···················· | ···················· | ····················· | ···················· | ···················· | ···················· | |
| M | 8283 | hCoV-19/England/ALDP-9FD89C/2020\|EPI_ISL_588023\|20 | ···················· | ···················· | ····················· | ···················· | ···················· | ···················· | |
| M | 8284 | hCoV-19/England/ALDP-9FDF1F/2020\|EPI_ISL_588024\|20 | ···················· | ···················· | ····················· | ···················· | ···················· | ···················· | |
| M | 8285 | hCoV-19/England/ALDP-9FCEC8/2020\|EPI_ISL_588025\|20 | ···················· | ···················· | ····················· | ···················· | ···················· | ···················· | |
| M | 8286 | hCoV-19/England/ALDP-9FD164/2020\|EPI_ISL_588027\|20 | ···················· | ···················· | ····················· | ···················· | ···················· | ···················· | |
| M | 8287 | hCoV-19/England/ALDP-9EE37F/2020\|EPI_ISL_589280\|20 | ···················· | ···················A | ····················· | ···················· | ···················· | ···················· | 1 |
| M | 8288 | hCoV-19/England/ALDP-9ED5E3/2020\|EPI_ISL_589281\|20 | ···················· | ···················· | ····················· | ···················· | ···················· | ···················· | |
| M | 8289 | hCoV-19/England/ALDP-9E56E8/2020\|EPI_ISL_589282\|20 | ···················· | ···················· | ····················· | ···················· | ···················· | ···················· | |
| M | 8290 | hCoV-19/England/ALDP-9ECDA0/2020\|EPI_ISL_589283\|20 | ···················· | ···················· | ····················· | ···················· | ···················· | ···················· | |

| | | | | | | | |
|---|---|---|---|---|---|---|---|
| M | 8291 hCoV-19/England/ALDP-9EDDBE/2020\|EPI_ISL_589284\|20 | .................... | .................... | .................... | .................... | .................... | .................... |
| M | 8292 hCoV-19/England/ALDP-9ED817/2020\|EPI_ISL_589285\|20 | .................... | .................... | .................... | .................... | .................... | .................... |
| M | 8293 hCoV-19/England/ALDP-9ED01F/2020\|EPI_ISL_589286\|20 | .................... | .................... | .................... | .................... | ............T....... | .................... |
| M | 8294 hCoV-19/England/ALDP-9ECFB9/2020\|EPI_ISL_589287\|20 | .................... | .................... | .................... | .................... | .................... | .................... |
| M | 8295 hCoV-19/England/ALDP-9ED49B/2020\|EPI_ISL_589288\|20 | .................... | .................... | .................... | .................... | .................... | .................... |
| M | 8296 hCoV-19/England/ALDP-9EDF12/2020\|EPI_ISL_589290\|20 | .................... | .A.................. | .................... | .................... | .................... | .................... |
| M | 8297 hCoV-19/England/ALDP-9E5A46/2020\|EPI_ISL_589291\|20 | .................... | .................... | .................... | .................... | .................... | .................... |
| M | 8298 hCoV-19/England/ALDP-9E5C8C/2020\|EPI_ISL_589292\|20 | .................... | .A.................. | .................... | .................... | .................... | ...A................ |
| M | 8299 hCoV-19/England/ALDP-9E56CA/2020\|EPI_ISL_589295\|20 | .................... | .................... | .................... | .................... | .................... | .................... |
| M | 8300 hCoV-19/England/ALDP-9ED1EF/2020\|EPI_ISL_589296\|20 | .................... | .................... | .................... | .................... | .................... | .................... |
| M | 8301 hCoV-19/England/MILK-9ECC0D/2020\|EPI_ISL_589297\|20 | .................... | .................... | .................... | .................... | .................... | .................... |
| M | 8302 hCoV-19/England/ALDP-9E5CAA/2020\|EPI_ISL_589298\|20 | .................... | .................... | .................... | .................... | .................... | .................... |
| M | 8303 hCoV-19/England/ALDP-9ED5C5/2020\|EPI_ISL_589299\|20 | .................... | .................... | .................... | .................... | .................... | .................... |
| M | 8304 hCoV-19/England/ALDP-9ED04C/2020\|EPI_ISL_589340\|20 | .................... | .................... | .................... | .................... | .................... | .................... |
| M | 8305 hCoV-19/England/ALDP-9EE39D/2020\|EPI_ISL_589346\|20 | .................... | .................... | .................... | .................... | .................... | .................... |
| M | 8306 hCoV-19/England/ALDP-9EE315/2020\|EPI_ISL_589347\|20 | .................... | .................... | .................... | .................... | .................... | .................... |
| M | 8307 hCoV-19/England/ALDP-9E5879/2020\|EPI_ISL_589349\|20 | .................... | .................... | .................... | .................... | .................... | .................... |
| M | 8308 hCoV-19/England/ALDP-9ED510/2020\|EPI_ISL_589350\|20 | .................... | .................... | .................... | .................... | .................... | .................... |
| M | 8309 hCoV-19/England/ALDP-9EDFD6/2020\|EPI_ISL_589352\|20 | .................... | .................... | .................... | .................... | .................... | .................... |
| M | 8310 hCoV-19/England/ALDP-9EE157/2020\|EPI_ISL_589353\|20 | .................... | .................... | .................... | .................... | .................... | .................... |
| M | 8311 hCoV-19/England/ALDP-9E57C7/2020\|EPI_ISL_589354\|20 | .................... | .................... | .................... | .................... | .................... | .................... |
| M | 8312 hCoV-19/England/MILK-9ECCB2/2020\|EPI_ISL_589355\|20 | .................... | .................... | .................... | .................... | .................... | .................... |
| M | 8313 hCoV-19/England/ALDP-9ECFC8/2020\|EPI_ISL_589356\|20 | .................... | .................... | .................... | .................... | .................... | .................... |
| M | 8314 hCoV-19/England/ALDP-9E576D/2020\|EPI_ISL_589357\|20 | .................... | .................... | .................... | .................... | .................... | .................... |
| M | 8315 hCoV-19/England/ALDP-9EE38E/2020\|EPI_ISL_589358\|20 | .................... | .................... | .................... | .................... | .................... | .................... |
| M | 8316 hCoV-19/England/ALDP-9E58E2/2020\|EPI_ISL_589359\|20 | .................... | .................... | .................... | .................... | .................... | .................... |
| M | 8317 hCoV-19/England/ALDP-9EE218/2020\|EPI_ISL_589400\|20 | .................... | .................... | .................... | .................... | .................... | .................... |
| M | 8318 hCoV-19/England/ALDP-9ECEE9/2020\|EPI_ISL_589401\|20 | .................... | .................... | .................... | .................... | .................... | .................... |
| M | 8319 hCoV-19/England/ALDP-9ED47D/2020\|EPI_ISL_589402\|20 | .................... | .................... | .................... | .................... | .................... | .................... |
| M | 8320 hCoV-19/England/ALDP-9ED853/2020\|EPI_ISL_589403\|20 | .................... | .................... | .................... | .................... | .................... | .................... |
| M | 8321 hCoV-19/England/ALDP-9ED352/2020\|EPI_ISL_589405\|20 | .................... | .................... | .................... | .................... | .................... | .................... |
| M | 8322 hCoV-19/England/ALDP-9ED334/2020\|EPI_ISL_589406\|20 | .................... | .................... | .................... | .................... | .................... | .................... |
| M | 8323 hCoV-19/England/MILK-9ECC85/2020\|EPI_ISL_589407\|20 | .................... | .................... | .................... | .................... | .................... | .................... |
| M | 8324 hCoV-19/England/ALDP-9EDF4F/2020\|EPI_ISL_589408\|20 | .................... | .................... | .................... | .................... | .................... | .................... |
| M | 8325 hCoV-19/England/ALDP-9EE306/2020\|EPI_ISL_589409\|20 | .................... | .................... | .................... | .................... | .................... | .................... |
| M | 8326 hCoV-19/England/ALDP-9EE1C0/2020\|EPI_ISL_589410\|20 | .................... | .................... | .................... | .................... | .................... | .................... |
| M | 8327 hCoV-19/England/ALDP-9ECDDD/2020\|EPI_ISL_589411\|20 | .................... | .................... | .................... | .................... | .................... | .................... |
| M | 8328 hCoV-19/England/ALDP-9E579A/2020\|EPI_ISL_589412\|20 | .................... | .................... | .................... | .................... | .................... | .................... |
| M | 8329 hCoV-19/England/ALDP-9E5CC8/2020\|EPI_ISL_589413\|20 | .................... | .................... | .................... | .................... | .................... | .................... |
| M | 8330 hCoV-19/England/ALDP-9E58F1/2020\|EPI_ISL_589414\|20 | .................... | .................... | .................... | .................... | .................... | .................... |
| M | 8331 hCoV-19/England/QEUH-9B4E71/2020\|EPI_ISL_590262\|20 | .................... | .................... | .................... | .................... | .................... | .................... |
| M | 8332 hCoV-19/England/ALDP-9B59AC/2020\|EPI_ISL_590263\|20 | .................... | .A.................. | .................... | .................... | .................... | ...A................ |
| M | 8333 hCoV-19/England/QEUH-9B50B6/2020\|EPI_ISL_590264\|20 | .................... | .................... | .................... | .................... | .................... | .................... |
| M | 8334 hCoV-19/England/ALDP-9B5B00/2020\|EPI_ISL_590265\|20 | .................... | .................... | .................... | .................... | .................... | .................... |
| M | 8335 hCoV-19/England/QEUH-9B4E9F/2020\|EPI_ISL_590266\|20 | .................... | .................... | .................... | .................... | .................... | .................... |
| M | 8336 hCoV-19/England/QEUH-9B556C/2020\|EPI_ISL_590267\|20 | .................... | ....A............... | .................... | .................... | .................... | .................... |
| M | 8337 hCoV-19/England/QEUH-9B4DDE/2020\|EPI_ISL_590268\|20 | .................... | .................... | .................... | .................... | ....T............... | .................... |
| M | 8338 hCoV-19/England/QEUH-9B465B/2020\|EPI_ISL_590269\|20 | .................... | .................... | .................... | .................... | .................... | .................... |
| M | 8339 hCoV-19/England/QEUH-9B564B/2020\|EPI_ISL_590270\|20 | .................... | .................... | .................... | .................... | .................... | .................... |
| M | 8340 hCoV-19/NorthernIreland/QEUH-9B5335/2020\|EPI_ISL_5 | .................... | .................... | .................... | .................... | .................... | .................... |
| M | 8341 hCoV-19/England/ALDP-9B5942/2020\|EPI_ISL_590272\|20 | .................... | .................... | .................... | .................... | .................... | .................... |
| M | 8342 hCoV-19/England/QEUH-9B4BC5/2020\|EPI_ISL_590273\|20 | .................... | .................... | .................... | .................... | .................... | .................... |
| M | 8343 hCoV-19/England/ALDP-9B5B4C/2020\|EPI_ISL_590274\|20 | .................... | .................... | .................... | .................... | .................... | .................... |
| M | 8344 hCoV-19/England/ALDP-9B5BE2/2020\|EPI_ISL_590275\|20 | .................... | .................... | .................... | .................... | .................... | .................... |
| M | 8345 hCoV-19/England/ALDP-9B57C0/2020\|EPI_ISL_590276\|20 | .................... | .................... | .................... | .................... | .................... | .................... |
| M | 8346 hCoV-19/England/ALDP-9B5AF4/2020\|EPI_ISL_590277\|20 | .................... | .................... | .................... | .................... | .................... | .................... |
| M | 8347 hCoV-19/England/QEUH-9B4882/2020\|EPI_ISL_590278\|20 | .................... | .................... | .................... | .................... | .................... | .................... |
| M | 8348 hCoV-19/England/QEUH-9B464C/2020\|EPI_ISL_590279\|20 | .................... | .................... | .................... | .................... | .................... | .................... |
| M | 8349 hCoV-19/England/QEUH-9B4C77/2020\|EPI_ISL_590280\|20 | .................... | .................... | .................... | .................... | .................... | .................... |
| M | 8350 hCoV-19/England/QEUH-9B4A04/2020\|EPI_ISL_590281\|20 | .................... | .................... | .................... | .................... | .................... | .................... |
| M | 8351 hCoV-19/England/ALDP-9B5B3D/2020\|EPI_ISL_590282\|20 | .................... | .................... | .................... | .................... | .................... | .................... |
| M | 8352 hCoV-19/England/ALDP-9B59E8/2020\|EPI_ISL_590283\|20 | .................... | .A.................. | .................... | .................... | .................... | ...A................ |
| M | 8353 hCoV-19/England/QEUH-9B4873/2020\|EPI_ISL_590320\|20 | .................... | .................... | .................... | .................... | .................... | .................... |
| M | 8354 hCoV-19/England/QEUH-9B55F3/2020\|EPI_ISL_590325\|20 | .................... | .................... | .................... | .................... | .................... | .................... |
| M | 8355 hCoV-19/England/ALDP-9B5A9A/2020\|EPI_ISL_590327\|20 | .................... | .................... | .................... | .................... | .................... | .................... |
| M | 8356 hCoV-19/England/ALDP-9B59F7/2020\|EPI_ISL_590328\|20 | .................... | .A.................. | .................... | .................... | .................... | .................... |
| M | 8357 hCoV-19/England/QEUH-9B4CB3/2020\|EPI_ISL_590330\|20 | .................... | .A.................. | .................... | .................... | .................... | ...A................ |
| M | 8358 hCoV-19/England/QEUH-9B53AE/2020\|EPI_ISL_590331\|20 | .................... | .................... | .................... | .................... | .................... | .................... |
| M | 8359 hCoV-19/England/QEUH-9B51A4/2020\|EPI_ISL_590332\|20 | .................... | .................... | .................... | .................... | .................... | .................... |
| M | 8360 hCoV-19/England/QEUH-9B5283/2020\|EPI_ISL_590333\|20 | .................... | .................... | .................... | .................... | .................... | .................... |
| M | 8361 hCoV-19/England/QEUH-9B4D29/2020\|EPI_ISL_590334\|20 | .................... | .................... | .................... | .................... | .................... | .................... |
| M | 8362 hCoV-19/England/ALDP-9B5B2E/2020\|EPI_ISL_590335\|20 | .................... | .................... | .................... | .................... | .................... | .................... |
| M | 8363 hCoV-19/England/QEUH-9B4837/2020\|EPI_ISL_590336\|20 | .................... | .................... | .................... | .................... | .................... | .................... |
| M | 8364 hCoV-19/England/ALDP-9B5924/2020\|EPI_ISL_590337\|20 | .................... | .................... | .................... | .................... | .................... | .................... |
| M | 8365 hCoV-19/England/QEUH-9B4E17/2020\|EPI_ISL_590338\|20 | .................... | .................... | .................... | .................... | .................... | .................... |
| M | 8366 hCoV-19/England/QEUH-9B49CB/2020\|EPI_ISL_590339\|20 | .................... | .................... | .................... | .................... | .................... | .................... |
| M | 8367 hCoV-19/Australia/VIC14313/2020\|EPI_ISL_591718\|202 | .................... | .................... | .................... | .................... | .................... | .................... |
| M | 8369 hCoV-19/Australia/VIC14746/2020\|EPI_ISL_592982\|202 | .................... | .................... | .................... | .................... | .................... | .................... |
| M | 8370 hCoV-19/Australia/VIC14757/2020\|EPI_ISL_592987\|202 | .................... | .................... | .................... | .................... | .................... | .................... |
| M | 8371 hCoV-19/Australia/VIC14763/2020\|EPI_ISL_592988\|202 | .................... | .................... | .................... | .................... | .................... | .................... |

| | | | | | | | |
|---|---|---|---|---|---|---|---|
| M | 8372 hCoV-19/Australia/VIC14764/2020\|EPI_ISL_592989\|202 | ···················· | ···················· | ···················· | ···················· | ···················· | ···················· |
| M | 8373 hCoV-19/Australia/VIC14767/2020\|EPI_ISL_592990\|202 | ···················· | ···················· | ···················· | ···················· | ···················· | ···················· |
| M | 8374 hCoV-19/Australia/VIC14768/2020\|EPI_ISL_592991\|202 | ···················· | ···················· | ···················· | ···················· | ···················· | ···················· |
| M | 8375 hCoV-19/Australia/VIC14770/2020\|EPI_ISL_592992\|202 | ···················· | ···················· | ···················· | ···················· | ···················· | ···················· |
| M | 8376 hCoV-19/Australia/VIC14775/2020\|EPI_ISL_592993\|202 | ···················· | ···················· | ···················· | ···················· | ···················· | ···················· |
| M | 8377 hCoV-19/Australia/VIC14776/2020\|EPI_ISL_592994\|202 | ···················· | ···················· | ···················· | ···················· | ···················· | ···················· |
| M | 8378 hCoV-19/Australia/VIC14778/2020\|EPI_ISL_592995\|202 | ···················· | ···················· | ···················· | ···················· | ···················· | ···················· |
| M | 8380 hCoV-19/Australia/VIC14782/2020\|EPI_ISL_592997\|202 | ···················· | ···················· | ···················· | ···················· | ···················· | ···················· |
| M | 8381 hCoV-19/Australia/VIC14786/2020\|EPI_ISL_592998\|202 | ···················· | ···················· | ···················· | ···················· | ···················· | ···················· |
| M | 8382 hCoV-19/Australia/VIC14787/2020\|EPI_ISL_592999\|202 | ···················· | ···················· | ···················· | ···················· | ···················· | ···················· |
| M | 8383 hCoV-19/England/NORW-F0BD5/2020\|EPI_ISL_595201\|202 | ···················· | ···················· | ···················· | ···················· | ···················· | ···················· |
| M | 8384 hCoV-19/England/NORW-F0BF3/2020\|EPI_ISL_595203\|202 | ···················· | ···················· | ···················· | ···················· | ···················· | ···················· |
| M | 8385 hCoV-19/England/NORW-F0C1E/2020\|EPI_ISL_595205\|202 | ···················· | ···················· | ···················· | ···················· | ···················· | ···················· |
| M | 8386 hCoV-19/England/NORW-F0C2D/2020\|EPI_ISL_595206\|202 | ···················· | ···················· | ···················· | ···················· | ···················· | ···················· |
| M | 8387 hCoV-19/England/NORW-F0C69/2020\|EPI_ISL_595210\|202 | ···················· | ···················· | ···················· | ···················· | ···················· | ···················· |
| M | 8390 hCoV-19/England/NORW-F11D2/2020\|EPI_ISL_595219\|202 | ···················· | ···················· | ···················· | ···················· | ···················· | ···················· |
| M | 8391 hCoV-19/England/NORW-F1275/2020\|EPI_ISL_595222\|202 | ···················· | ···················· | ···················· | ···················· | ···················· | ···················· |
| M | 8392 hCoV-19/England/NORW-F1309/2020\|EPI_ISL_595224\|202 | ···················· | ···················· | ···················· | ···················· | ···················· | ···················· |
| M | 8394 hCoV-19/England/NORW-F1433/2020\|EPI_ISL_595228\|202 | ···················· | ···················· | ···················· | ···················· | ···················· | ···················· |
| M | 8397 hCoV-19/England/NOTT-114B70/2020\|EPI_ISL_595232\|20 | ···················· | ···················· | ···················· | ···················· | ···················· | ···················· |
| M | 8398 hCoV-19/England/NOTT-114BF8/2020\|EPI_ISL_595233\|20 | ···················· | ···················· | ···················· | ···················· | ···················· | ···················· |
| M | 8399 hCoV-19/England/NOTT-114C40/2020\|EPI_ISL_595235\|20 | ···················· | ···················· | ···················· | ···················· | ···················· | ···················· |
| M | 8400 hCoV-19/England/NOTT-114F1A/2020\|EPI_ISL_595236\|20 | ···················· | ···················· | ···················· | ···················· | ···················· | ···················· |
| M | 8401 hCoV-19/England/NOTT-115043/2020\|EPI_ISL_595237\|20 | ···················· | ···················· | ···················· | ···················· | ···················· | ···················· |
| M | 8402 hCoV-19/England/NOTT-1153A4/2020\|EPI_ISL_595239\|20 | ···················· | ···················· | ···················· | ···················· | ···················· | ···················· |
| M | 8403 hCoV-19/England/NOTT-1153C2/2020\|EPI_ISL_595240\|20 | ···················· | ···················· | ···················· | ···················· | ···················· | ···················· |
| M | 8404 hCoV-19/England/NOTT-1153D1/2020\|EPI_ISL_595241\|20 | ···················· | ···················· | ···················· | ···················· | ···················· | ···················· |
| M | 8405 hCoV-19/England/NOTT-1153E0/2020\|EPI_ISL_595242\|20 | ···················· | ···················· | ···················· | ···················· | ···················· | ···················· |
| M | 8406 hCoV-19/England/NOTT-1153FF/2020\|EPI_ISL_595243\|20 | ···················· | ···················· | ···················· | ···················· | ···················· | ···················· |
| M | 8407 hCoV-19/England/NOTT-115492/2020\|EPI_ISL_595244\|20 | ···················· | ···················· | ···················· | ···················· | ···················· | ···················· |
| M | 8408 hCoV-19/England/NOTT-115508/2020\|EPI_ISL_595245\|20 | ···················· | ···················· | ···················· | ···················· | ···················· | ···················· |
| M | 8409 hCoV-19/England/NOTT-115580/2020\|EPI_ISL_595247\|20 | ···················· | ······A············· | ···················· | ···················· | ···················· | ···················· |
| M | 8410 hCoV-19/England/NOTT-11559F/2020\|EPI_ISL_595248\|20 | ···················· | ···················· | ···················· | ···················· | ···················· | ···················· |
| M | 8411 hCoV-19/England/NOTT-1155AE/2020\|EPI_ISL_595249\|20 | ···················· | ···················· | ···················· | ···················· | ···················· | ···················· |
| M | 8412 hCoV-19/England/NOTT-1155CC/2020\|EPI_ISL_595251\|20 | ···················· | ·A·················· | ···················· | ···················· | ···················· | ···A················ |
| M | 8413 hCoV-19/England/NOTT-115632/2020\|EPI_ISL_595252\|20 | ···················· | ···················· | ···················· | ···················· | ···················· | ···················· |
| M | 8414 hCoV-19/England/NOTT-115641/2020\|EPI_ISL_595253\|20 | ···················· | ···················· | ···················· | ···················· | ···················· | ···················· |
| M | 8415 hCoV-19/England/NOTT-115650/2020\|EPI_ISL_595254\|20 | ···················· | ···················· | ···················· | ···················· | ···················· | ···················· |
| M | 8416 hCoV-19/England/NOTT-11567E/2020\|EPI_ISL_595255\|20 | ···················· | ···················· | ···················· | ···················· | ···················· | ···················· |
| M | 8417 hCoV-19/England/NOTT-11575D/2020\|EPI_ISL_595258\|20 | ···················· | ···················· | ···················· | ···················· | ···················· | ···················· |
| M | 8418 hCoV-19/England/NOTT-11576C/2020\|EPI_ISL_595259\|20 | ···················· | ···················· | ···················· | ···················· | ···················· | ···················· |
| M | 8419 hCoV-19/England/NOTT-11577B/2020\|EPI_ISL_595260\|20 | ···················· | ···················· | ···················· | ···················· | ···················· | ···················· |
| M | 8420 hCoV-19/England/NOTT-11578A/2020\|EPI_ISL_595261\|20 | ···················· | ···················· | ···················· | ···················· | ···················· | ···················· |
| M | 8421 hCoV-19/England/NOTT-115799/2020\|EPI_ISL_595262\|20 | ···················· | ···················· | ···················· | ···················· | ···················· | ···················· |
| M | 8422 hCoV-19/England/NOTT-1157A8/2020\|EPI_ISL_595263\|20 | ···················· | ···················· | ···················· | ···················· | ···················· | ···················· |
| M | 8423 hCoV-19/England/NOTT-1157C6/2020\|EPI_ISL_595264\|20 | ···················· | ···················· | ···················· | ···················· | ···················· | ···················· |
| M | 8424 hCoV-19/England/NOTT-1157D5/2020\|EPI_ISL_595265\|20 | ···················· | ···················· | ···················· | ···················· | ···················· | ···················· |
| M | 8425 hCoV-19/England/NOTT-11581E/2020\|EPI_ISL_595267\|20 | ···················· | ···················· | ···················· | ···················· | ···················· | ···················· |
| M | 8426 hCoV-19/England/NOTT-11583C/2020\|EPI_ISL_595269\|20 | ···················· | ···················· | ···················· | ···················· | ···················· | ···················· |
| M | 8427 hCoV-19/England/NOTT-11584B/2020\|EPI_ISL_595270\|20 | ···················· | ···················· | ···················· | ···················· | ···················· | ···················· |
| M | 8428 hCoV-19/England/NOTT-115887/2020\|EPI_ISL_595271\|20 | ···················· | ···················· | ···················· | ···················· | ···················· | ···················· |
| M | 8429 hCoV-19/England/NOTT-1158E1/2020\|EPI_ISL_595272\|20 | ···················· | ···················· | ···················· | ···················· | ···················· | ···················· |
| M | 8430 hCoV-19/England/NOTT-115939/2020\|EPI_ISL_595274\|20 | ···················· | ···················· | ···················· | ···················· | ···················· | ···················· |
| M | 8431 hCoV-19/England/NOTT-115984/2020\|EPI_ISL_595275\|20 | ···················· | ···················· | ···················· | ···················· | ···················· | ···················· |
| M | 8432 hCoV-19/England/NOTT-1159C0/2020\|EPI_ISL_595278\|20 | ···················· | ···················· | ···················· | ···················· | ···················· | ···················· |
| M | 8433 hCoV-19/England/NOTT-1159DF/2020\|EPI_ISL_595279\|20 | ···················· | ···················· | ···················· | ···················· | ···················· | ···················· |
| M | 8434 hCoV-19/England/NOTT-115A27/2020\|EPI_ISL_595280\|20 | ···················· | ···················· | ···················· | ···················· | ···················· | ···················· |
| M | 8435 hCoV-19/England/NOTT-115A36/2020\|EPI_ISL_595281\|20 | ···················· | ···················· | ···················· | ···················· | ···················· | ···················· |
| M | 8436 hCoV-19/England/NOTT-115A54/2020\|EPI_ISL_595282\|20 | ···················· | ···················· | ···················· | ···················· | ···················· | ···················· |
| M | 8437 hCoV-19/England/NOTT-115ACD/2020\|EPI_ISL_595283\|20 | ···················· | ···················· | ···················· | ···················· | ···················· | ···················· |
| M | 8438 hCoV-19/England/NOTT-115ADC/2020\|EPI_ISL_595284\|20 | ···················· | ···················· | ···················· | ···················· | ···················· | ···················· |
| M | 8439 hCoV-19/England/NOTT-115B06/2020\|EPI_ISL_595285\|20 | ···················· | ···················· | ···················· | ···················· | ···················· | ···················· |
| M | 8440 hCoV-19/England/NOTT-115B42/2020\|EPI_ISL_595286\|20 | ···················· | ···················· | ···················· | ···················· | ···················· | ···················· |
| M | 8441 hCoV-19/England/NOTT-115CB8/2020\|EPI_ISL_595289\|20 | ···················· | ···················· | ···················· | ···················· | ···················· | ···················· |
| M | 8442 hCoV-19/England/NOTT-115CC7/2020\|EPI_ISL_595290\|20 | ···················· | ···················· | ···················· | ···················· | ···················· | ···················· |
| M | 8443 hCoV-19/England/NOTT-115CE5/2020\|EPI_ISL_595291\|20 | ···················· | ···················· | ···················· | ···················· | ···················· | ···················· |
| M | 8444 hCoV-19/England/NOTT-115D5B/2020\|EPI_ISL_595292\|20 | ···················· | ···················· | ···················· | ···················· | ···················· | ···················· |
| M | 8445 hCoV-19/England/NOTT-115D6A/2020\|EPI_ISL_595293\|20 | ···················· | ···················· | ···················· | ···················· | ···················· | ···················· |
| M | 8446 hCoV-19/England/NOTT-115D79/2020\|EPI_ISL_595294\|20 | ···················· | ···················· | ···················· | ···················· | ···················· | ···················· |
| M | 8447 hCoV-19/England/NOTT-115F64/2020\|EPI_ISL_595296\|20 | ···················· | ···················· | ···················· | ···················· | ···················· | ···················· |
| M | 8448 hCoV-19/England/NOTT-115F73/2020\|EPI_ISL_595297\|20 | ···················· | ···················· | ···················· | ···················· | ···················· | ···················· |
| M | 8450 hCoV-19/Wales/PHWC-16E315/2020\|EPI_ISL_585701\|2020 | ···················· | ···················· | ···················· | ···················· | ···················· | ···················· |
| M | 8452 hCoV-19/Wales/PHWC-16E351/2020\|EPI_ISL_585704\|2020 | ···················· | ···················· | ···················· | ···················· | ···················· | ···················· |
| M | 8454 hCoV-19/Wales/PHWC-16E37F/2020\|EPI_ISL_585706\|2020 | ···················· | ···················· | ···················· | ···················· | ···················· | ···················· |
| M | 8455 hCoV-19/Wales/PHWC-16E38E/2020\|EPI_ISL_585707\|2020 | ···················· | ···················· | ···················· | ···················· | ···················· | ···················· |
| M | 8456 hCoV-19/Wales/PHWC-16E3AC/2020\|EPI_ISL_585708\|2020 | ···················· | ···················· | ···················· | ···················· | ···················· | ···················· |
| M | 8459 hCoV-19/Wales/PHWC-16E3D9/2020\|EPI_ISL_585711\|2020 | ···················· | ···················· | ···················· | ···················· | ···················· | ···················· |
| M | 8461 hCoV-19/Wales/PHWC-16E3F7/2020\|EPI_ISL_585713\|2020 | ···················· | ···················· | ···················· | ···················· | ···················· | ···················· |
| M | 8462 hCoV-19/Wales/PHWC-16E45E/2020\|EPI_ISL_585718\|2020 | ···················· | ···················· | ···················· | ···················· | ···················· | ···················· |
| M | 8466 hCoV-19/Wales/PHWC-16E49A/2020\|EPI_ISL_585722\|2020 | ···················· | ···················· | ···················· | ···················· | ···················· | ···················· |

| | | | | | | | |
|---|---|---|---|---|---|---|---|
| M | 8468 hCoV-19/Wales/PHWC-16E4B8/2020\|EPI_ISL_585724\|2020 | .................... | .................... | .................... | .................... | .................... | .................... |
| M | 8470 hCoV-19/Wales/PHWC-16E4E5/2020\|EPI_ISL_585727\|2020 | .................... | .................... | .................... | .................... | .................... | .................... |
| M | 8471 hCoV-19/Wales/MILK-9F240C/2020\|EPI_ISL_589200\|2020 | .................... | .................... | .................... | .................... | .................... | .................... |
| M | 8472 hCoV-19/England/QEUH-9F955D/2020\|EPI_ISL_589201\|20 | .................... | .................... | .................... | .................... | .................... | .................... |
| M | 8473 hCoV-19/England/QEUH-9F75C8/2020\|EPI_ISL_589202\|20 | .................... | .................... | .................... | .................... | .................... | .................... |
| M | 8474 hCoV-19/England/QEUH-9F762F/2020\|EPI_ISL_589203\|20 | .................... | .................... | .................... | .................... | .................... | .................... |
| M | 8475 hCoV-19/England/QEUH-9F7BF3/2020\|EPI_ISL_589204\|20 | .................... | .................... | .................... | .................... | .................... | .................... |
| M | 8476 hCoV-19/England/MILK-9F29EF/2020\|EPI_ISL_589205\|20 | .................... | .A.................. | .................... | .................... | .................... | .................... |
| M | 8477 hCoV-19/England/QEUH-9F9669/2020\|EPI_ISL_589206\|20 | .................... | .................... | .................... | .................... | .................... | .................... |
| M | 8478 hCoV-19/England/QEUH-9F75F5/2020\|EPI_ISL_589207\|20 | .................... | .................... | .................... | .................... | .................... | .................... |
| M | 8479 hCoV-19/England/QEUH-9F7B8A/2020\|EPI_ISL_589209\|20 | .................... | .................... | .................... | .................... | .................... | .................... |
| M | 8480 hCoV-19/England/QEUH-9F9A8B/2020\|EPI_ISL_589210\|20 | .................... | .................... | .................... | .................... | .................... | .................... |
| M | 8481 hCoV-19/England/MILK-9F275E/2020\|EPI_ISL_589211\|20 | .................... | .................... | .................... | .................... | .................... | .................... |
| M | 8482 hCoV-19/England/MILK-9F22F3/2020\|EPI_ISL_589212\|20 | .................... | .................... | .................... | .................... | .................... | .................... |
| M | 8483 hCoV-19/England/QEUH-9F7944/2020\|EPI_ISL_589213\|20 | .................... | .................... | .................... | .................... | .................... | .................... |
| M | 8484 hCoV-19/England/QEUH-9F75D7/2020\|EPI_ISL_589215\|20 | .................... | .................... | .................... | .................... | .................... | .................... |
| M | 8485 hCoV-19/England/QEUH-9F7B20/2020\|EPI_ISL_589216\|20 | .................... | .................... | .................... | .................... | .................... | .................... |
| M | 8486 hCoV-19/England/QEUH-9F953F/2020\|EPI_ISL_589219\|20 | .................... | .................... | .................... | .................... | .................... | .................... |
| M | 8487 hCoV-19/England/QEUH-9F9CB2/2020\|EPI_ISL_589220\|20 | .................... | .................... | .................... | .................... | .................... | .................... |
| M | 8488 hCoV-19/England/MILK-9F2518/2020\|EPI_ISL_589221\|20 | .................... | .................... | .................... | .................... | .................... | .................... |
| M | 8489 hCoV-19/England/QEUH-9F9757/2020\|EPI_ISL_589222\|20 | .................... | .................... | .................... | .................... | .................... | .................... |
| M | 8490 hCoV-19/England/QEUH-9F97A2/2020\|EPI_ISL_589223\|20 | .................... | .................... | .................... | .................... | .................... | .................... |
| M | 8491 hCoV-19/England/MILK-9F218D/2020\|EPI_ISL_589224\|20 | .................... | .................... | .................... | .................... | .................... | .................... |
| M | 8492 hCoV-19/England/MILK-9F2B34/2020\|EPI_ISL_589226\|20 | .................... | .................... | .................... | .................... | .................... | .................... |
| M | 8493 hCoV-19/England/MILK-9F2062/2020\|EPI_ISL_589227\|20 | .................... | .................... | .................... | .................... | .................... | .................... |
| M | 8494 hCoV-19/England/MILK-9F223F/2020\|EPI_ISL_589231\|20 | .................... | .................... | .................... | .................... | ......T............. | .................... |
| M | 8495 hCoV-19/England/QEUH-9F9881/2020\|EPI_ISL_589232\|20 | .................... | .................... | .................... | .................... | .................... | .................... |
| M | 8496 hCoV-19/England/MILK-9F2A55/2020\|EPI_ISL_589233\|20 | .................... | .................... | .................... | .................... | .................... | .................... |
| M | 8497 hCoV-19/England/QEUH-9F9511/2020\|EPI_ISL_589239\|20 | .................... | .................... | .................... | .................... | .................... | .................... |
| M | 8498 hCoV-19/England/QEUH-9F997F/2020\|EPI_ISL_589240\|20 | .................... | .................... | .................... | .................... | .................... | .................... |
| M | 8499 hCoV-19/England/QEUH-9F99E8/2020\|EPI_ISL_589241\|20 | .................... | .................... | .................... | .................... | .................... | .................... |
| M | 8500 hCoV-19/England/MILK-9F2B07/2020\|EPI_ISL_589242\|20 | .................... | .................... | .................... | .................... | .................... | .................... |
| M | 8501 hCoV-19/England/MILK-9F2800/2020\|EPI_ISL_589243\|20 | .................... | .................... | .................... | .................... | .................... | .................... |
| M | 8502 hCoV-19/England/QEUH-9F7E54/2020\|EPI_ISL_589244\|20 | .................... | .................... | .................... | .................... | .................... | .................... |
| M | 8503 hCoV-19/England/QEUH-9F7865/2020\|EPI_ISL_589245\|20 | .................... | .................... | .................... | .................... | .................... | .................... |
| M | 8504 hCoV-19/England/QEUH-9F9924/2020\|EPI_ISL_589246\|20 | .................... | .A.................. | .................... | .................... | .................... | .................... |
| M | 8505 hCoV-19/England/MILK-9F2730/2020\|EPI_ISL_589247\|20 | .................... | .................... | .................... | .................... | .................... | .................... |
| M | 8506 hCoV-19/England/QEUH-9F9793/2020\|EPI_ISL_589249\|20 | .................... | .................... | .................... | .................... | .................... | .................... |
| M | 8507 hCoV-19/England/QEUH-9F7A7E/2020\|EPI_ISL_589250\|20 | .................... | .................... | .................... | .................... | .................... | .................... |
| M | 8508 hCoV-19/England/MILK-9F2B25/2020\|EPI_ISL_589251\|20 | .................... | .................... | .................... | .................... | .................... | .................... |
| M | 8509 hCoV-19/England/QEUH-9F96A5/2020\|EPI_ISL_589252\|20 | .................... | .................... | .................... | .................... | .................... | .................... |
| M | 8510 hCoV-19/England/QEUH-9F7AAB/2020\|EPI_ISL_589254\|20 | .................... | .................... | .................... | .................... | .................... | .................... |
| M | 8511 hCoV-19/England/QEUH-9F97EE/2020\|EPI_ISL_589255\|20 | .................... | .................... | .................... | .................... | .................... | .................... |
| M | 8512 hCoV-19/England/MILK-9F25EB/2020\|EPI_ISL_589256\|20 | .................... | .................... | .................... | .................... | .................... | .................... |
| M | 8513 hCoV-19/England/QEUH-9F7A41/2020\|EPI_ISL_589257\|20 | .................... | .................... | .................... | .................... | .................... | .................... |
| M | 8514 hCoV-19/England/QEUH-9F7B3F/2020\|EPI_ISL_589258\|20 | .................... | .................... | .................... | .................... | .................... | .................... |
| M | 8515 hCoV-19/Wales/MILK-9F2A91/2020\|EPI_ISL_589259\|2020 | .................... | .................... | .................... | .................... | .................... | .................... |
| M | 8516 hCoV-19/England/MILK-9F230F/2020\|EPI_ISL_589260\|20 | .................... | .................... | .................... | .................... | .................... | .................... |
| M | 8517 hCoV-19/England/MILK-9F2B16/2020\|EPI_ISL_589261\|20 | .................... | .................... | .................... | .................... | .................... | .................... |
| M | 8518 hCoV-19/England/QEUH-9F7C5A/2020\|EPI_ISL_589263\|20 | .................... | .................... | .................... | .................... | .................... | .................... |
| M | 8519 hCoV-19/England/MILK-9F233C/2020\|EPI_ISL_589265\|20 | .................... | .................... | .................... | .................... | .................... | .................... |
| M | 8520 hCoV-19/England/MILK-9F281F/2020\|EPI_ISL_589266\|20 | .................... | .................... | .................... | .................... | .................... | .................... |
| M | 8521 hCoV-19/England/ALDP-9ED589/2020\|EPI_ISL_589268\|20 | .................... | .................... | .................... | .................... | .................... | .................... |
| M | 8522 hCoV-19/England/ALDP-9E5800/2020\|EPI_ISL_589269\|20 | .................... | .................... | .................... | .................... | .................... | .................... |
| M | 8523 hCoV-19/England/ALDP-9E5888/2020\|EPI_ISL_589270\|20 | .................... | .................... | .................... | .................... | .................... | .................... |
| M | 8524 hCoV-19/England/ALDP-9ECECB/2020\|EPI_ISL_589271\|20 | .................... | .................... | .................... | .................... | .................... | .................... |
| M | 8525 hCoV-19/England/ALDP-9ED370/2020\|EPI_ISL_589272\|20 | .................... | .A.................. | .................... | .................... | .................... | .................... |
| M | 8526 hCoV-19/England/ALDP-9ECEF8/2020\|EPI_ISL_589273\|20 | .................... | .................... | .................... | .................... | .................... | .................... |
| M | 8527 hCoV-19/England/ALDP-9ED273/2020\|EPI_ISL_589274\|20 | .................... | .................... | .................... | .................... | .................... | .................... |
| M | 8528 hCoV-19/England/ALDP-9EE227/2020\|EPI_ISL_589275\|20 | .................... | .................... | .................... | .................... | .................... | .................... |
| M | 8529 hCoV-19/England/ALDP-9E58A6/2020\|EPI_ISL_589276\|20 | .................... | .................... | .................... | .................... | .................... | .................... |
| M | 8530 hCoV-19/England/ALDP-9ED185/2020\|EPI_ISL_589277\|20 | .................... | .................... | .................... | .................... | .................... | .................... |
| M | 8531 hCoV-19/England/ALDP-9E582E/2020\|EPI_ISL_589278\|20 | .................... | .................... | .................... | .................... | .................... | .................... |
| M | 8532 hCoV-19/England/ALDP-9EDF21/2020\|EPI_ISL_589279\|20 | .................... | .................... | .................... | .................... | .................... | .................... |
| M | 8533 hCoV-19/England/ALDP-9E59B2/2020\|EPI_ISL_589294\|20 | .................... | .................... | .................... | .................... | .................... | .................... |
| M | 8534 hCoV-19/England/ALDP-9B5A30/2020\|EPI_ISL_590200\|20 | .................... | .................... | .................... | .................... | .................... | .................... |
| M | 8535 hCoV-19/England/QEUH-9B463D/2020\|EPI_ISL_590201\|20 | .................... | .................... | .................... | .................... | .................... | .................... |
| M | 8536 hCoV-19/England/QEUH-9B52B0/2020\|EPI_ISL_590202\|20 | .................... | .................... | .................... | .................... | .................... | .................... |
| M | 8537 hCoV-19/England/QEUH-9B560F/2020\|EPI_ISL_590203\|20 | .................... | .................... | .................... | .................... | .................... | .................... |
| M | 8538 hCoV-19/England/QEUH-9B4934/2020\|EPI_ISL_590204\|20 | .................... | .................... | .................... | .................... | .................... | .................... |
| M | 8539 hCoV-19/England/QEUH-9B48DD/2020\|EPI_ISL_590205\|20 | .................... | .................... | .................... | .................... | .................... | .................... |
| M | 8540 hCoV-19/England/ALDP-9B5AD6/2020\|EPI_ISL_590206\|20 | .................... | .................... | .................... | .................... | .................... | .................... |
| M | 8541 hCoV-19/England/QEUH-9B52FC/2020\|EPI_ISL_590207\|20 | .................... | .................... | .................... | .................... | .................... | .................... |
| M | 8542 hCoV-19/England/QEUH-9B498F/2020\|EPI_ISL_590208\|20 | .................... | .................... | .................... | .................... | .................... | .................... |
| M | 8543 hCoV-19/England/QEUH-9B502F/2020\|EPI_ISL_590209\|20 | .................... | .................... | .................... | .................... | .................... | .................... |
| M | 8544 hCoV-19/England/QEUH-9B4C68/2020\|EPI_ISL_590211\|20 | .................... | .................... | .................... | .................... | .................... | .................... |
| M | 8545 hCoV-19/England/QEUH-9B4DA1/2020\|EPI_ISL_590212\|20 | .................... | .................... | .................... | .................... | ....T............... | .................... |
| M | 8546 hCoV-19/England/ALDP-9B5C76/2020\|EPI_ISL_590213\|20 | .................... | .................... | .................... | .................... | .................... | .................... |
| M | 8547 hCoV-19/England/ALDP-9B5C58/2020\|EPI_ISL_590214\|20 | .................... | .................... | .................... | .................... | .................... | .................... |
| M | 8548 hCoV-19/England/ALDP-9B5A5E/2020\|EPI_ISL_590220\|20 | .................... | .................... | .................... | .................... | .................... | .................... |

```
M 8549 hCoV-19/England/QEUH-9B49E9/2020|EPI_ISL_590221|20 ···················· ···················· ···················· ···················· ···················· ····················
M 8550 hCoV-19/England/QEUH-9B5353/2020|EPI_ISL_590222|20 ···················· ···················· ···················· ···················· ···················· ····················
M 8551 hCoV-19/England/QEUH-9B4776/2020|EPI_ISL_590223|20 ···················· ···················· ···················· ···················· ···················· ····················
M 8552 hCoV-19/England/ALDP-9B57EE/2020|EPI_ISL_590224|20 ···················· ···················· ···················· ···················· ···················· ····················
M 8553 hCoV-19/England/ALDP-9B5757/2020|EPI_ISL_590225|20 ···················· ···················· ···················· ···················· ···················· ····················
M 8554 hCoV-19/England/ALDP-9B5AB8/2020|EPI_ISL_590226|20 ···················· ···················· ···················· ···················· ···················· ····················
M 8555 hCoV-19/England/QEUH-9B54E7/2020|EPI_ISL_590227|20 ···················· ···················· ···················· ···················· ···················· ····················
M 8556 hCoV-19/England/ALDP-9B5C67/2020|EPI_ISL_590228|20 ···················· ···················· ···················· ···················· ···················· ····················
M 8557 hCoV-19/England/QEUH-9B46C4/2020|EPI_ISL_590229|20 ···················· ·A·················· ···················· ···················· ···················· ··A·················
M 8558 hCoV-19/England/QEUH-9B5010/2020|EPI_ISL_590230|20 ···················· ···················· ···················· ···················· ···················· ····················
M 8559 hCoV-19/England/QEUH-9B4679/2020|EPI_ISL_590231|20 ···················· ···················· ···················· ···················· ···················· ····················
M 8560 hCoV-19/England/QEUH-9B55D5/2020|EPI_ISL_590232|20 ···················· ···················· ···················· ···················· ···················· ····················
M 8561 hCoV-19/England/QEUH-9B4F50/2020|EPI_ISL_590233|20 ···················· ···················· ···················· ···················· ···················· ····················
M 8562 hCoV-19/England/ALDP-9B58CD/2020|EPI_ISL_590234|20 ···················· ···················· ···················· ···················· ···················· ····················
M 8563 hCoV-19/England/ALDP-9B5B97/2020|EPI_ISL_590235|20 ···················· ···················· ···················· ···················· ···················· ····················
M 8564 hCoV-19/England/QEUH-9B56C3/2020|EPI_ISL_590237|20 ···················· ···················· ···················· ···················· ···················· ····················
M 8565 hCoV-19/England/QEUH-9B503E/2020|EPI_ISL_590238|20 ···················· ···················· ···················· ···················· ······T············· ····················
M 8566 hCoV-19/England/ALDP-9B5BB5/2020|EPI_ISL_590239|20 ···················· ···················· ···················· ···················· ···················· ····················
M 8567 hCoV-19/England/QEUH-9B51C2/2020|EPI_ISL_590240|20 ···················· ···················· ···················· ···················· ···················· ····················
M 8568 hCoV-19/England/QEUH-9B4A9B/2020|EPI_ISL_590241|20 ···················· ···················· ···················· ···················· ···················· ····················
M 8569 hCoV-19/England/QEUH-9B4952/2020|EPI_ISL_590242|20 ···················· ···················· ···················· ···················· ···················· ····················
M 8570 hCoV-19/England/QEUH-9B48BF/2020|EPI_ISL_590243|20 ···················· ···················· ···················· ···················· ···················· ····················
M 8571 hCoV-19/England/QEUH-9B5256/2020|EPI_ISL_590244|20 ···················· ···················· ···················· ···················· ···················· ····················
M 8572 hCoV-19/England/QEUH-9B4B4D/2020|EPI_ISL_590245|20 ···················· ···················· ···················· ···················· ···················· ····················
M 8573 hCoV-19/England/ALDP-9B58AF/2020|EPI_ISL_590246|20 ···················· ···················· ···················· ···················· ···················· ····················
M 8574 hCoV-19/England/QEUH-9B4970/2020|EPI_ISL_590247|20 ···················· ···················· ···················· ···················· ···················· ····················
M 8575 hCoV-19/England/QEUH-9B4AB9/2020|EPI_ISL_590248|20 ···················· ···················· ···················· ···················· ···················· ····················
M 8576 hCoV-19/England/QEUH-9B4F41/2020|EPI_ISL_590249|20 ···················· ···················· ···················· ···················· ···················· ····················
M 8577 hCoV-19/England/ALDP-9B5766/2020|EPI_ISL_590250|20 ···················· ·A·················· ···················· ···················· ···················· ····················
M 8578 hCoV-19/England/QEUH-9B48CE/2020|EPI_ISL_590251|20 ···················· ···················· ···················· ···················· ···················· ····················
M 8579 hCoV-19/England/QEUH-9B4D0B/2020|EPI_ISL_590252|20 ···················· ···················· ···················· ···················· ···················· ····················
M 8580 hCoV-19/England/QEUH-9B5274/2020|EPI_ISL_590253|20 ···················· ···················· ···················· ···················· ···················· ····················
M 8581 hCoV-19/England/QEUH-9B47C1/2020|EPI_ISL_590254|20 ···················· ···················· ···················· ···················· ···················· ····················
M 8582 hCoV-19/England/ALDP-9B57FD/2020|EPI_ISL_590255|20 ···················· ···················· ···················· ···················· ···················· ····················
M 8583 hCoV-19/England/QEUH-9B4FBA/2020|EPI_ISL_590256|20 ···················· ······C············· ···················· ···················· ···················· ····················
M 8584 hCoV-19/England/ALDP-9B5A7C/2020|EPI_ISL_590257|20 ···················· ···················· ···················· ···················· ···················· ····················
M 8585 hCoV-19/England/QEUH-9B48EC/2020|EPI_ISL_590258|20 ···················· ···················· ···················· ···················· ···················· ····················
M 8586 hCoV-19/England/ALDP-9B5BD3/2020|EPI_ISL_590260|20 ···················· ···················· ···················· ···················· ···················· ····················
M 8587 hCoV-19/England/ALDP-9B5915/2020|EPI_ISL_590284|20 ···················· ···················· ···················· ···················· ···················· ····················
M 8588 hCoV-19/England/QEUH-9B55B7/2020|EPI_ISL_590285|20 ···················· ···················· ···················· ···················· ···················· ····················
M 8589 hCoV-19/England/ALDP-9B5863/2020|EPI_ISL_590286|20 ···················· ·A·················· ···················· ···················· ···················· ····················
M 8590 hCoV-19/England/QEUH-9B53BD/2020|EPI_ISL_590289|20 ···················· ···················· ···················· ···················· ···················· ····················
M 8591 hCoV-19/England/QEUH-9B462E/2020|EPI_ISL_590290|20 ···················· ···················· ···················· ···················· ······T············· ····················
M 8592 hCoV-19/England/QEUH-9B4E80/2020|EPI_ISL_590291|20 ···················· ···················· ···················· ···················· ···················· ····················
M 8593 hCoV-19/England/QEUH-9B4B5C/2020|EPI_ISL_590292|20 ···················· ···················· ···················· ···················· ···················· ····················
M 8594 hCoV-19/England/QEUH-9B4891/2020|EPI_ISL_590293|20 ···················· ···················· ···················· ···················· ···················· ····················
M 8595 hCoV-19/England/QEUH-9B5229/2020|EPI_ISL_590294|20 ···················· ·A·················· ···················· ···················· ···················· ··A·················
M 8596 hCoV-19/England/ALDP-9B5827/2020|EPI_ISL_590295|20 ···················· ···················· ···················· ···················· ···················· ····················
M 8597 hCoV-19/England/ALDP-9B5739/2020|EPI_ISL_590296|20 ···················· ···················· ···················· ···················· ···················· ····················
M 8598 hCoV-19/England/QEUH-9B539F/2020|EPI_ISL_590297|20 ···················· ···················· ···················· ···················· ······T············· ····················
M 8599 hCoV-19/England/QEUH-9B4828/2020|EPI_ISL_590298|20 ···················· ···················· ···················· ···················· ···················· ····················
M 8600 hCoV-19/England/QEUH-9B5362/2020|EPI_ISL_590299|20 ···················· ···················· ···················· ···················· ···················· ····················
M 8601 hCoV-19/Australia/NSW1004/2020|EPI_ISL_591500|2020 ···················· ···················· ···················· ···················· ···················· ····················
M 8602 hCoV-19/Australia/NSW1005/2020|EPI_ISL_591501|2020 ···················· ···················· ···················· ···················· ···················· ····················
M 8603 hCoV-19/Australia/NSW989/2020|EPI_ISL_591502|2020- ···················· ···················· ···················· ···················· ···················· ····················
M 8604 hCoV-19/Australia/NSW992/2020|EPI_ISL_591503|2020- ···················· ···················· ···················· ···················· ···················· ····················
M 8605 hCoV-19/Australia/NSW993/2020|EPI_ISL_591504|2020- ···················· ···················· ···················· ···················· ···················· ····················
M 8606 hCoV-19/Australia/NSW1001/2020|EPI_ISL_591505|2020 ···················· ···················· ···················· ···················· ···················· ····················
M 8607 hCoV-19/Australia/NSW1002/2020|EPI_ISL_591506|2020 ···················· ···················· ···················· ···················· ···················· ····················
M 8608 hCoV-19/Australia/NSW986/2020|EPI_ISL_591507|2020- ········T··········· ···················· ···················· ···················· ···················· ····················
M 8609 hCoV-19/Australia/NSW980/2020|EPI_ISL_591508|2020- ···················· ···················· ···················· ···················· ···················· ····················
M 8610 hCoV-19/Australia/NSW983/2020|EPI_ISL_591509|2020- ···················· ···················· ···················· ···················· ···················· ····················
M 8611 hCoV-19/Australia/NSW994/2020|EPI_ISL_591510|2020- ···················· ···················· ···················· ···················· ···················· ····················
M 8612 hCoV-19/Australia/NSW999/2020|EPI_ISL_591511|2020- ···················· ···················· ···················· ···················· ···················· ····················
M 8613 hCoV-19/Australia/NSW1000/2020|EPI_ISL_591512|2020 ···················· ···················· ···················· ···················· ···················· ····················
M 8614 hCoV-19/Australia/NSW1006/2020|EPI_ISL_591513|2020 ···················· ···················· ···················· ···················· ···················· ····················
M 8615 hCoV-19/Australia/NSW1007/2020|EPI_ISL_591514|2020 ···················· ···················· ···················· ···················· ···················· ····················
M 8616 hCoV-19/Australia/NSW1008/2020|EPI_ISL_591515|2020 ···················· ···················· ···················· ···················· ···················· ····················
M 8617 hCoV-19/Australia/NSW1013/2020|EPI_ISL_591516|2020 ···················· ···················· ···················· ···················· ···················· ····················
M 8618 hCoV-19/Australia/NSW990/2020|EPI_ISL_591517|2020- ········T··········· ···················· ···················· ···················· ···················· ····················
M 8619 hCoV-19/Australia/NSW984/2020|EPI_ISL_591518|2020- ···················· ···················· ···················· ···················· ···················· ····················
M 8620 hCoV-19/Chile/RM-CMM-LRF3924/2020|EPI_ISL_591522|2 ···················· ···················· ···················· ···················· ···················· ····················
M 8621 hCoV-19/Chile/RM-CMM-NA4204/2020|EPI_ISL_591523|20 ···················· ···················· ···················· ···················· ···················· ····················
M 8622 hCoV-19/Chile/RM-CMM-HRG4342/2020|EPI_ISL_591526|2 ···················· ···················· ···················· ···················· ···················· ····················
M 8623 hCoV-19/Chile/RM-CMM-LAE7918/2020|EPI_ISL_591527|2 ···················· ···················· ···················· ···················· ···················· ····················
M 8624 hCoV-19/Chile/RM-CMM-ADC1858/2020|EPI_ISL_591528|2 ···················· ···················· ···················· ···················· ···················· ····················
M 8625 hCoV-19/Chile/RM-CMM-MCD9529/2020|EPI_ISL_591529|2 ···················· ···················· ···················· ···················· ···················· ····················
M 8626 hCoV-19/Chile/RM-CMM-EDB244K/2020|EPI_ISL_591530|2 ···················· ···················· ···················· ···················· ···················· ····················
M 8627 hCoV-19/Chile/RM-CMM-IN233883987/2020|EPI_ISL_5915 ···················· ···················· ···················· ···················· ···················· ····················
M 8628 hCoV-19/Chile/RM-CMM-IN533883977/2020|EPI_ISL_5915 ···················· ···················· ···················· ···················· ···················· ····················
```

| | | | | | | | |
|---|---|---|---|---|---|---|---|
| M | 8629 hCoV-19/Chile/RM-CMM-A2P533884024/2020\|EPI_ISL_591 | ···················· | ···················· | ····················· | ···················· | ···················· | ···················· |
| M | 8630 hCoV-19/Chile/RM-CMM-A5P533884002/2020\|EPI_ISL_591 | ···················· | ···················· | ····················· | ···················· | ···················· | ···················· |
| M | 8631 hCoV-19/Japan/IC-0013/2020\|EPI_ISL_591535\|2020-03 | ···················· | ···················· | ····················· | ···················· | ···················· | ···················· |
| M | 8632 hCoV-19/Japan/IC-0022/2020\|EPI_ISL_591536\|2020-04 | ···················· | ···················· | ····················· | ···················· | ···················· | ···················· |
| M | 8633 hCoV-19/Japan/IC-0031/2020\|EPI_ISL_591537\|2020-06 | ···················· | ···················· | ····················· | ···················· | ···················· | ···················· |
| M | 8634 hCoV-19/Japan/IC-0032/2020\|EPI_ISL_591538\|2020-06 | ···················· | ···················· | ····················· | ···················· | ···················· | ···················· |
| M | 8635 hCoV-19/Japan/IC-0090/2020\|EPI_ISL_591539\|2020-07 | ···················· | ···················· | ····················· | ···················· | ···················· | ···················· |
| M | 8636 hCoV-19/France/OCC-34/2020\|EPI_ISL_591542\|2020-06- | ···················· | ···················· | ····················· | ···················· | ···················· | ···················· |
| M | 8637 hCoV-19/France/OCC-35/2020\|EPI_ISL_591543\|2020-06- | ···················· | ···················· | ····················· | ···················· | ···················· | ···················· |
| M | 8638 hCoV-19/France/OCC-36/2020\|EPI_ISL_591544\|2020-06- | ···················· | ···················· | ····················· | ···················· | ···················· | ···················· |
| M | 8639 hCoV-19/France/OCC-37/2020\|EPI_ISL_591545\|2020-06- | ···················· | ···················· | ····················· | ···················· | ···················· | ···················· |
| M | 8640 hCoV-19/Australia/VIC15335/2020\|EPI_ISL_592820\|202 | ···················· | ···················· | ····················· | ···················· | ···················· | ···················· |
| M | 8641 hCoV-19/Australia/VIC15338/2020\|EPI_ISL_592821\|202 | ···················· | ···················· | ····················· | ···················· | ···················· | ···················· |
| M | 8642 hCoV-19/Australia/VIC14572/2020\|EPI_ISL_592822\|202 | ···················· | ···················· | ····················· | ···················· | ···················· | ···················· |
| M | 8643 hCoV-19/Australia/VIC14598/2020\|EPI_ISL_592823\|202 | ···················· | ···················· | ····················· | ···················· | ···················· | ···················· |
| M | 8644 hCoV-19/Australia/VIC14726/2020\|EPI_ISL_592824\|202 | ···················· | ···················· | ····················· | ···················· | ···················· | ···················· |
| M | 8645 hCoV-19/Australia/VIC14752/2020\|EPI_ISL_592825\|202 | ···················· | ···················· | ····················· | ···················· | ···················· | ···················· |
| M | 8646 hCoV-19/Australia/VIC14759/2020\|EPI_ISL_592826\|202 | ···················· | ···················· | ····················· | ···················· | ············R······· | ···················· |
| M | 8649 hCoV-19/Australia/VIC15098/2020\|EPI_ISL_592829\|202 | ···················· | ···················· | ····················· | ···················· | ···················· | ···················· |
| M | 8650 hCoV-19/Australia/VIC15115/2020\|EPI_ISL_592830\|202 | ···················· | ···················· | ····················· | ···················· | ···················· | ···················· |
| M | 8651 hCoV-19/Australia/VIC15151/2020\|EPI_ISL_592831\|202 | ···················· | ···················· | ····················· | ···················· | ···················· | ···················· |
| M | 8653 hCoV-19/Australia/VIC14541/2020\|EPI_ISL_592833\|202 | ···················· | ···················· | ····················· | ···················· | ···················· | ···················· |
| M | 8654 hCoV-19/Australia/VIC14560/2020\|EPI_ISL_592834\|202 | ···················· | ···················· | ····················· | ···················· | ···················· | ···················· |
| M | 8655 hCoV-19/Australia/VIC14614/2020\|EPI_ISL_592835\|202 | ···················· | ···················· | ····················· | ···················· | ···················· | ···················· |
| M | 8656 hCoV-19/Australia/VIC14630/2020\|EPI_ISL_592836\|202 | ···················· | ···················· | ····················· | ···················· | ···················· | ···················· |
| M | 8657 hCoV-19/Australia/VIC14659/2020\|EPI_ISL_592837\|202 | ···················· | ···················· | ····················· | ···················· | ···················· | ···················· |
| M | 8658 hCoV-19/Australia/VIC14668/2020\|EPI_ISL_592838\|202 | ···················· | ···················· | ····················· | ···················· | ···················· | ···················· |
| M | 8659 hCoV-19/Australia/VIC14705/2020\|EPI_ISL_592839\|202 | ···················· | ···················· | ····················· | ···················· | ···················· | ···················· |
| M | 8660 hCoV-19/Australia/VIC14858/2020\|EPI_ISL_592840\|202 | ···················· | ···················· | ····················· | ···················· | ···················· | ···················· |
| M | 8662 hCoV-19/Australia/VIC14946/2020\|EPI_ISL_592842\|202 | ···················· | ···················· | ····················· | ···················· | ···················· | ···················· |
| M | 8663 hCoV-19/Australia/VIC15106/2020\|EPI_ISL_592843\|202 | ···················· | ···················· | ····················· | ···················· | ···················· | ···················· |
| M | 8664 hCoV-19/Australia/VIC15116/2020\|EPI_ISL_592844\|202 | ···················· | ···················· | ····················· | ···················· | ···················· | ···················· |
| M | 8665 hCoV-19/Australia/VIC15169/2020\|EPI_ISL_592845\|202 | ···················· | ···················· | ····················· | ···················· | ···················· | ···················· |
| M | 8666 hCoV-19/Australia/VIC15175/2020\|EPI_ISL_592846\|202 | ···················· | ···················· | ····················· | ···················· | ···················· | ···················· |
| M | 8667 hCoV-19/Australia/VIC15201/2020\|EPI_ISL_592847\|202 | ···················· | ···················· | ····················· | ···················· | ···················· | ···················· |
| M | 8668 hCoV-19/Australia/VIC15247/2020\|EPI_ISL_592848\|202 | ···················· | ···················· | ····················· | ···················· | ···················· | ···················· |
| M | 8669 hCoV-19/Australia/VIC15262/2020\|EPI_ISL_592849\|202 | ···················· | ···················· | ····················· | ···················· | ···················· | ···················· |
| M | 8670 hCoV-19/Australia/VIC15346/2020\|EPI_ISL_592850\|202 | ···················· | ···················· | ····················· | ···················· | ···················· | ···················· |
| M | 8671 hCoV-19/Australia/VIC15352/2020\|EPI_ISL_592851\|202 | ···················· | ···················· | ····················· | ···················· | ···················· | ···················· |
| M | 8672 hCoV-19/Australia/VIC15426/2020\|EPI_ISL_592852\|202 | ···················· | ···················· | ····················· | ···················· | ···················· | ···················· |
| M | 8673 hCoV-19/Australia/VIC15436/2020\|EPI_ISL_592853\|202 | ···················· | ···················· | ····················· | ···················· | ···················· | ···················· |
| M | 8676 hCoV-19/Australia/VIC15810/2020\|EPI_ISL_592856\|202 | ···················· | ···················· | ····················· | ···················· | ···················· | ···················· |
| M | 8677 hCoV-19/Australia/VIC15252/2020\|EPI_ISL_592866\|202 | ···················· | ···················· | ····················· | ···················· | ···················· | ···················· |
| M | 8678 hCoV-19/Australia/VIC14511/2020\|EPI_ISL_592867\|202 | ···················· | ···················· | ····················· | ···················· | ···················· | ···················· |
| M | 8680 hCoV-19/Australia/VIC14513/2020\|EPI_ISL_592869\|202 | ···················· | ···················· | ····················· | ···················· | ···················· | ···················· |
| M | 8681 hCoV-19/Australia/VIC14517/2020\|EPI_ISL_592873\|202 | ···················· | ···················· | ····················· | ···················· | ···················· | ···················· |
| M | 8682 hCoV-19/Australia/VIC14520/2020\|EPI_ISL_592874\|202 | ···················· | ···················· | ····················· | ···················· | ···················· | ···················· |
| M | 8683 hCoV-19/Australia/VIC14522/2020\|EPI_ISL_592875\|202 | ···················· | ···················· | ····················· | ···················· | ···················· | ···················· |
| M | 8685 hCoV-19/Australia/VIC14532/2020\|EPI_ISL_592880\|202 | ···················· | ···················· | ··K·················· | ···················· | ···················· | ···················· |
| M | 8686 hCoV-19/Australia/VIC14536/2020\|EPI_ISL_592881\|202 | ···················· | ···················· | ··K·················· | ···················· | ···················· | ···················· |
| M | 8687 hCoV-19/Australia/VIC14540/2020\|EPI_ISL_592882\|202 | ···················· | ···················· | ····················· | ···················· | N··················· | ···················· |
| M | 8689 hCoV-19/Australia/VIC14544/2020\|EPI_ISL_592884\|202 | ···················· | ···················· | ····················· | ···················· | ···················· | ···················· |
| M | 8695 hCoV-19/Australia/VIC14555/2020\|EPI_ISL_592890\|202 | ···················· | ···················· | ····················· | ···················· | ···················· | ···················· |
| M | 8696 hCoV-19/Australia/VIC14556/2020\|EPI_ISL_592891\|202 | ···················· | ···················· | ····················· | ···················· | ···················· | ···················· |
| M | 8698 hCoV-19/Australia/VIC14558/2020\|EPI_ISL_592893\|202 | ···················· | ···················· | ····················· | ···················· | ···················· | ···················· |
| M | 8699 hCoV-19/Australia/VIC14563/2020\|EPI_ISL_592894\|202 | ···················· | ···················· | ····················· | ···················· | ···················· | ···················· |
| M | 8700 hCoV-19/Australia/VIC14564/2020\|EPI_ISL_592895\|202 | ···················· | ···················· | ····················· | ···················· | ···················· | ···················· |
| M | 8702 hCoV-19/Australia/VIC14566/2020\|EPI_ISL_592897\|202 | ···················· | ···················· | ····················· | ···················· | ···················· | ···················· |
| M | 8703 hCoV-19/Australia/VIC14567/2020\|EPI_ISL_592898\|202 | ···················· | ···················· | ····················· | ···················· | N··················· | ···················· |
| M | 8704 hCoV-19/Australia/VIC14568/2020\|EPI_ISL_592899\|202 | ···················· | ···················· | ····················· | ···················· | ···················· | ···················· |
| M | 8705 hCoV-19/England/CAMC-9EBB7A/2020\|EPI_ISL_584401\|20 | ···················· | ·A·················· | ····················· | ···················· | ···················· | ··A················· |
| M | 8706 hCoV-19/England/CAMC-9EBB98/2020\|EPI_ISL_584402\|20 | ···················· | ···················· | ····················· | ···················· | ···················· | ···················· |
| M | 8707 hCoV-19/England/CAMC-9EBBD4/2020\|EPI_ISL_584404\|20 | ···················· | ···················· | ····················· | ···················· | ···················· | ···················· |
| M | 8708 hCoV-19/England/CAMC-9EBC1D/2020\|EPI_ISL_584406\|20 | ···················· | ···················· | ····················· | ···················· | ···················· | ···················· |
| M | 8709 hCoV-19/England/CAMC-9EBC3B/2020\|EPI_ISL_584407\|20 | ···················· | ···················· | ····················· | ···················· | ···················· | ···················· |
| M | 8710 hCoV-19/England/CAMC-9EBC68/2020\|EPI_ISL_584409\|20 | ···················· | ···················· | ····················· | ···················· | ···················· | ···················· |
| M | 8711 hCoV-19/England/CAMC-9EBC95/2020\|EPI_ISL_584410\|20 | ···················· | ·A·················· | ····················· | ···················· | ···················· | ··A················· |
| M | 8712 hCoV-19/England/CAMC-9EBCA4/2020\|EPI_ISL_584411\|20 | ···················· | ···················· | ····················· | ···················· | ···················· | ···················· |
| M | 8713 hCoV-19/England/CAMC-9EBCC2/2020\|EPI_ISL_584412\|20 | ···················· | ···················· | ····················· | ···················· | ···················· | ···················· |
| M | 8714 hCoV-19/England/CAMC-9EBCE0/2020\|EPI_ISL_584413\|20 | ···················· | ···················· | ····················· | ···················· | ···················· | ···················· |
| M | 8715 hCoV-19/England/CAMC-9EBD29/2020\|EPI_ISL_584414\|20 | ···················· | ···················· | ····················· | ···················· | ···················· | ···················· |
| M | 8716 hCoV-19/England/CAMC-9EBD38/2020\|EPI_ISL_584415\|20 | ···················· | ···················· | ····················· | ···················· | ···················· | ···················· |
| M | 8717 hCoV-19/England/CAMC-9EBD74/2020\|EPI_ISL_584418\|20 | ···················· | ···················· | ····················· | ···················· | ···················· | ···················· |
| M | 8718 hCoV-19/England/CAMC-9EBD83/2020\|EPI_ISL_584419\|20 | ···················· | ···················· | ····················· | ···················· | ···················· | ···················· |
| M | 8719 hCoV-19/England/CAMC-9EBD92/2020\|EPI_ISL_584420\|20 | ···················· | ···················· | ····················· | ···················· | ···················· | ···················· |
| M | 8720 hCoV-19/England/CAMC-9EBDA1/2020\|EPI_ISL_584421\|20 | ···················· | ···················· | ····················· | ···················· | ······T············· | ···················· |
| M | 8721 hCoV-19/England/CAMC-9EBDCF/2020\|EPI_ISL_584422\|20 | ···················· | ···················· | ····················· | ···················· | ···················· | ···················· |
| M | 8722 hCoV-19/England/CAMC-9EBDDE/2020\|EPI_ISL_584423\|20 | ···················· | ···················· | ····················· | ···················· | ···················· | ···················· |
| M | 8723 hCoV-19/England/CAMC-9EBE08/2020\|EPI_ISL_584424\|20 | ···················· | ···················· | ····················· | ···················· | ···················· | ···················· |
| M | 8724 hCoV-19/England/CAMC-9EBEBD/2020\|EPI_ISL_584425\|20 | ···················· | ···················· | ····················· | ···················· | ···················· | ···················· |

| | | | | | | | |
|---|---|---|---|---|---|---|---|
| M | 8725 hCoV-19/England/CAMC-9EBECC/2020\|EPI_ISL_584426\|20 | ···················· | ···················· | ···················· | ···················· | ···················· | ···················· |
| M | 8726 hCoV-19/England/CAMC-9EBEDB/2020\|EPI_ISL_584427\|20 | ···················· | ···················· | ···················· | ···················· | ···················· | ···················· |
| M | 8727 hCoV-19/England/CAMC-9EBEEA/2020\|EPI_ISL_584428\|20 | ···················· | ···················· | ···················· | ···················· | ···················· | ···················· |
| M | 8728 hCoV-19/England/CAMC-9EBEF9/2020\|EPI_ISL_584429\|20 | ···················· | ·A·················· | ···················· | ···················· | ···················· | ··A················· |
| M | 8729 hCoV-19/England/CAMC-9EBF05/2020\|EPI_ISL_584430\|20 | ···················· | ···················· | ···················· | ···················· | ···················· | ···················· |
| M | 8730 hCoV-19/England/CAMC-9EBF14/2020\|EPI_ISL_584431\|20 | ···················· | ···················· | ···················· | ···················· | ···················· | ···················· |
| M | 8731 hCoV-19/England/CAMC-9EBF50/2020\|EPI_ISL_584433\|20 | ···················· | ·A·················· | ···················· | ···················· | ···················· | ··A················· |
| M | 8732 hCoV-19/England/CAMC-9EBF7E/2020\|EPI_ISL_584434\|20 | ···················· | ·A·················· | ···················· | ···················· | ···················· | ··A················· |
| M | 8733 hCoV-19/England/CAMC-9EBFD8/2020\|EPI_ISL_584436\|20 | ···················· | ···················· | ···················· | ···················· | ···················· | ···················· |
| M | 8734 hCoV-19/England/CAMC-9EBFF6/2020\|EPI_ISL_584437\|20 | ···················· | ·A·················· | ···················· | ···················· | ···················· | ···················· |
| M | 8735 hCoV-19/England/CAMC-9EC05C/2020\|EPI_ISL_584439\|20 | ···················· | ·A·················· | ···················· | ···················· | ···················· | ···················· |
| M | 8736 hCoV-19/England/CWAR-24FD85F/2020\|EPI_ISL_584440\|2 | ···················· | ···················· | ···················· | ···················· | ···················· | ···················· |
| M | 8737 hCoV-19/England/CWAR-24FD8B9/2020\|EPI_ISL_584446\|2 | ···················· | ···················· | ···················· | ···················· | ···················· | ···················· |
| M | 8738 hCoV-19/England/CWAR-24FD9B6/2020\|EPI_ISL_584460\|2 | ···················· | ···················· | ···················· | ···················· | ···················· | ···················· |
| M | 8739 hCoV-19/England/CWAR-24FD9C5/2020\|EPI_ISL_584461\|2 | ···················· | ···················· | ···················· | ···················· | ···········T········ | ···················· |
| M | 8740 hCoV-19/England/CWAR-24FD9D4/2020\|EPI_ISL_584462\|2 | ···················· | ···················· | ···················· | ···················· | ···················· | ···················· |
| M | 8741 hCoV-19/England/CWAR-24FDA0E/2020\|EPI_ISL_584465\|2 | ···················· | ···················· | ···················· | ···················· | ···················· | ···················· |
| M | 8743 hCoV-19/England/CWAR-24FDA2C/2020\|EPI_ISL_584467\|2 | ···················· | ···················· | ···················· | ···················· | ···················· | ···················· |
| M | 8744 hCoV-19/England/CWAR-24FDA3B/2020\|EPI_ISL_584468\|2 | ···················· | ···················· | ···················· | ···················· | ···················· | ···················· |
| M | 8745 hCoV-19/England/CWAR-24FDA4A/2020\|EPI_ISL_584469\|2 | ···················· | ···················· | ···················· | ···················· | ···················· | ···················· |
| M | 8746 hCoV-19/England/CWAR-24FDA59/2020\|EPI_ISL_584470\|2 | ···················· | ···················· | ···················· | ···················· | ···················· | ···················· |
| M | 8747 hCoV-19/England/CWAR-24FDA68/2020\|EPI_ISL_584471\|2 | ···················· | ···················· | ···················· | ···················· | ···················· | ···················· |
| M | 8748 hCoV-19/England/CWAR-24FDA77/2020\|EPI_ISL_584472\|2 | ···················· | ···················· | ···················· | ···················· | ···················· | ···················· |
| M | 8749 hCoV-19/England/CWAR-24FDA86/2020\|EPI_ISL_584473\|2 | ···················· | ···················· | ···················· | ···················· | ···················· | ···················· |
| M | 8750 hCoV-19/England/CWAR-24FDA95/2020\|EPI_ISL_584474\|2 | ···················· | ···················· | ···················· | ···················· | ···················· | ···················· |
| M | 8751 hCoV-19/England/CWAR-24FDAA4/2020\|EPI_ISL_584475\|2 | ···················· | ···················· | ···················· | ···················· | ···················· | ···················· |
| M | 8753 hCoV-19/England/CWAR-24FDAE0/2020\|EPI_ISL_584478\|2 | ···················· | ···················· | ···················· | ···················· | ···················· | ···················· |
| M | 8754 hCoV-19/England/CWAR-24FDAFF/2020\|EPI_ISL_584479\|2 | ···················· | ···················· | ···················· | ···················· | ···················· | ···················· |
| M | 8755 hCoV-19/England/CWAR-24FDB0B/2020\|EPI_ISL_584480\|2 | ···················· | ···················· | ···················· | ···················· | ···················· | ···················· |
| M | 8756 hCoV-19/England/CWAR-24FDB1A/2020\|EPI_ISL_584481\|2 | ···················· | ···················· | ···················· | ···················· | ···················· | ···················· |
| M | 8757 hCoV-19/England/CWAR-24FDB29/2020\|EPI_ISL_584482\|2 | ···················· | ···················· | ···················· | ···················· | ···················· | ···················· |
| M | 8758 hCoV-19/England/CWAR-24FDB56/2020\|EPI_ISL_584485\|2 | ···················· | ···················· | ···················· | ···················· | ···················· | ···················· |
| M | 8759 hCoV-19/England/CWAR-24FDB83/2020\|EPI_ISL_584488\|2 | ···················· | ···················· | ···················· | ···················· | ···················· | ···················· |
| M | 8760 hCoV-19/England/CWAR-24FDB92/2020\|EPI_ISL_584489\|2 | ···················· | ···················· | ···················· | ···················· | ···················· | ···················· |
| M | 8761 hCoV-19/England/CWAR-24FDBA1/2020\|EPI_ISL_584490\|2 | ···················· | ···················· | ···················· | ···················· | ···················· | ···················· |
| M | 8762 hCoV-19/England/CWAR-24FDBB0/2020\|EPI_ISL_584491\|2 | ···················· | ···················· | ···················· | ···················· | ···················· | ···················· |
| M | 8763 hCoV-19/England/CWAR-24FDBCF/2020\|EPI_ISL_584492\|2 | ···················· | ···················· | ···················· | ···················· | ···················· | ···················· |
| M | 8764 hCoV-19/England/CWAR-24FDBDE/2020\|EPI_ISL_584493\|2 | ···················· | ···················· | ···················· | ···················· | ···················· | ···················· |
| M | 8765 hCoV-19/England/CWAR-24FDC08/2020\|EPI_ISL_584496\|2 | ···················· | ···················· | ···················· | ···················· | ···················· | ···················· |
| M | 8766 hCoV-19/England/CWAR-24FDC17/2020\|EPI_ISL_584497\|2 | ···················· | ···················· | ···················· | ···················· | ···················· | ···················· |
| M | 8767 hCoV-19/England/CWAR-24FDC26/2020\|EPI_ISL_584498\|2 | ···················· | ···················· | ···················· | ···················· | ···················· | ···················· |
| M | 8768 hCoV-19/Wales/PHWC-16E51F/2020\|EPI_ISL_585730\|2020 | ···················· | ···················· | ···················· | ···················· | ···················· | ···················· |
| M | 8769 hCoV-19/Wales/PHWC-16E52E/2020\|EPI_ISL_585731\|2020 | ···················· | ···················· | ···················· | ···················· | ···················· | ···················· |
| M | 8771 hCoV-19/Wales/PHWC-16E56A/2020\|EPI_ISL_585734\|2020 | ···················· | ···················· | ···················· | ···················· | ···················· | ···················· |
| M | 8772 hCoV-19/Wales/PHWC-16E579/2020\|EPI_ISL_585735\|2020 | ···················· | ···················· | ···················· | ···················· | ···················· | ···················· |
| M | 8774 hCoV-19/Wales/PHWC-16E597/2020\|EPI_ISL_585737\|2020 | ···················· | ···················· | ···················· | ···················· | ···················· | ···················· |
| M | 8776 hCoV-19/Wales/PHWC-16E5C4/2020\|EPI_ISL_585740\|2020 | ···················· | ···················· | ···················· | ···················· | ···················· | ···················· |
| M | 8778 hCoV-19/Wales/PHWC-16E5E2/2020\|EPI_ISL_585742\|2020 | ···················· | ···················· | ···················· | ···················· | ···················· | ···················· |
| M | 8779 hCoV-19/Wales/PHWC-16E5F1/2020\|EPI_ISL_585743\|2020 | ···················· | ···················· | ···················· | ···················· | ···················· | ···················· |
| M | 8780 hCoV-19/Wales/PHWC-16E62B/2020\|EPI_ISL_585745\|2020 | ···················· | ···················· | ···················· | ···················· | ·····T·············· | ···················· |
| M | 8781 hCoV-19/Wales/PHWC-16E63A/2020\|EPI_ISL_585746\|2020 | ···················· | ···················· | ···················· | ···················· | ···················· | ···················· |
| M | 8786 hCoV-19/Wales/PHWC-16E6A3/2020\|EPI_ISL_585753\|2020 | ···················· | ···················· | ···················· | ···················· | ···················· | ···················· |
| M | 8787 hCoV-19/Wales/PHWC-16E6C1/2020\|EPI_ISL_585755\|2020 | ···················· | ···················· | ···················· | ···················· | ···················· | ···················· |
| M | 8791 hCoV-19/Wales/PHWC-16E737/2020\|EPI_ISL_585762\|2020 | ···················· | ···················· | ···················· | ···················· | ···················· | ···················· |
| M | 8793 hCoV-19/Wales/PHWC-16E791/2020\|EPI_ISL_585767\|2020 | ···················· | ···················· | ···················· | ···················· | ···················· | ···················· |
| M | 8794 hCoV-19/Wales/PHWC-16E7BF/2020\|EPI_ISL_585768\|2020 | ···················· | ···················· | ···················· | ···················· | ···················· | ···················· |
| M | 8795 hCoV-19/Wales/PHWC-16E7DD/2020\|EPI_ISL_585770\|2020 | ···················· | ···················· | ···················· | ···················· | ···················· | ···················· |
| M | 8796 hCoV-19/Wales/PHWC-16E7EC/2020\|EPI_ISL_585771\|2020 | ···················· | ···················· | ···················· | ···················· | ···················· | ···················· |
| M | 8797 hCoV-19/Wales/PHWC-16E7FB/2020\|EPI_ISL_585772\|2020 | ···················· | ···················· | ···················· | ···················· | ···················· | ···················· |
| M | 8798 hCoV-19/Wales/PHWC-16E816/2020\|EPI_ISL_585774\|2020 | ···················· | ···················· | ···················· | ···················· | ···················· | ···················· |
| M | 8800 hCoV-19/Wales/PHWC-16E834/2020\|EPI_ISL_585776\|2020 | ···················· | ···················· | ···················· | ···················· | ···················· | ···················· |
| M | 8802 hCoV-19/Wales/PHWC-16E852/2020\|EPI_ISL_585778\|2020 | ···················· | ···················· | ···················· | ···················· | ···················· | ···················· |
| M | 8803 hCoV-19/Wales/PHWC-16E861/2020\|EPI_ISL_585779\|2020 | ···················· | ···················· | ···················· | ···················· | ···················· | ···················· |
| M | 8804 hCoV-19/Wales/PHWC-16E870/2020\|EPI_ISL_585780\|2020 | ···················· | ···················· | ···················· | ···················· | ···················· | ···················· |
| M | 8805 hCoV-19/Wales/PHWC-16E89E/2020\|EPI_ISL_585782\|2020 | ···················· | ···················· | ···················· | ···················· | ···················· | ···················· |
| M | 8806 hCoV-19/Wales/PHWC-16E8AD/2020\|EPI_ISL_585783\|2020 | ···················· | ···················· | ···················· | ···················· | ···················· | ···················· |
| M | 8807 hCoV-19/Wales/PHWC-16E8BC/2020\|EPI_ISL_585784\|2020 | ···················· | ···················· | ···················· | ···················· | ···················· | ···················· |
| M | 8808 hCoV-19/Wales/PHWC-16E8CB/2020\|EPI_ISL_585785\|2020 | ···················· | ···················· | ···················· | ···················· | ···················· | ···················· |
| M | 8809 hCoV-19/Wales/PHWC-16E8DA/2020\|EPI_ISL_585786\|2020 | ···················· | ···················· | ···················· | ···················· | ···················· | ···················· |
| M | 8810 hCoV-19/Wales/PHWC-16E8E9/2020\|EPI_ISL_585787\|2020 | ···················· | ···················· | ···················· | ···················· | ···················· | ···················· |
| M | 8811 hCoV-19/Wales/PHWC-16E8F8/2020\|EPI_ISL_585788\|2020 | ···················· | ···················· | ···················· | ···················· | ···················· | ···················· |
| M | 8812 hCoV-19/Wales/PHWC-16E904/2020\|EPI_ISL_585789\|2020 | ···················· | ···················· | ···················· | ···················· | ···················· | ···················· |
| M | 8814 hCoV-19/Wales/PHWC-16E922/2020\|EPI_ISL_585791\|2020 | ···················· | ···················· | ···················· | ···················· | ···················· | ···················· |
| M | 8815 hCoV-19/Wales/PHWC-16E931/2020\|EPI_ISL_585792\|2020 | ···················· | ···················· | ···················· | ···················· | ···················· | ···················· |
| M | 8816 hCoV-19/Wales/PHWC-16E940/2020\|EPI_ISL_585793\|2020 | ···················· | ···················· | ···················· | ···················· | ···················· | ···················· |
| M | 8817 hCoV-19/Wales/PHWC-16E95F/2020\|EPI_ISL_585794\|2020 | ···················· | ···················· | ···················· | ···················· | ···················· | ···················· |
| M | 8818 hCoV-19/Wales/PHWC-16E96E/2020\|EPI_ISL_585795\|2020 | ···················· | ···················· | ···················· | ···················· | ···················· | ···················· |
| M | 8819 hCoV-19/Wales/PHWC-16E98C/2020\|EPI_ISL_585797\|2020 | ···················· | ···················· | ···················· | ···················· | ···················· | ···················· |
| M | 8820 hCoV-19/Wales/PHWC-16E99B/2020\|EPI_ISL_585798\|2020 | ···················· | ···················· | ···················· | ···················· | ···················· | ···················· |
| M | 8821 hCoV-19/Wales/PHWC-16E9AA/2020\|EPI_ISL_585799\|2020 | ···················· | ···················· | ···················· | ···················· | ···················· | ···················· |

| | | | | | | | | |
|---|---|---|---|---|---|---|---|---|
| M | 8822 hCoV-19/England/MILK-9F285B/2020\|EPI_ISL_589160\|20 | ···················· | ···················· | ···················· | ···················· | ···················· | ···················· | |
| M | 8823 hCoV-19/Wales/MILK-9F2A0A/2020\|EPI_ISL_589161\|2020 | ···················· | ···················· | ···················· | ···················· | ···················· | ···················· | |
| M | 8824 hCoV-19/England/QEUH-9F7AF6/2020\|EPI_ISL_589162\|20 | ···················· | ·A·················· | ···················· | ···················· | ···················· | ···················· | |
| M | 8825 hCoV-19/England/QEUH-9F7A6F/2020\|EPI_ISL_589163\|20 | ···················· | ···················· | ···················· | ···················· | ···················· | ···················· | |
| M | 8826 hCoV-19/England/QEUH-9F960F/2020\|EPI_ISL_589164\|20 | ···················· | ···················· | ···················· | ···················· | ···················· | ···················· | |
| M | 8827 hCoV-19/England/QEUH-9F7953/2020\|EPI_ISL_589165\|20 | ···················· | ···················· | ···················· | ···················· | ···················· | ···················· | |
| M | 8828 hCoV-19/England/MILK-9F2202/2020\|EPI_ISL_589166\|20 | ···················· | ···················· | ···················· | ···················· | ···················· | ···················· | |
| M | 8829 hCoV-19/England/MILK-9F2572/2020\|EPI_ISL_589167\|20 | ···················· | ···················· | ···················· | ···················· | ···················· | ···················· | |
| M | 8830 hCoV-19/England/MILK-9F2633/2020\|EPI_ISL_589168\|20 | ···················· | ···················· | ···················· | ···················· | ···················· | ···················· | |
| M | 8831 hCoV-19/England/MILK-9F2475/2020\|EPI_ISL_589169\|20 | ···················· | ···················· | ···················· | ···················· | ···················· | ···················· | |
| M | 8832 hCoV-19/England/MILK-9F2493/2020\|EPI_ISL_589235\|20 | ···················· | ···················· | ···················· | ···················· | ···················· | ···················· | |
| M | 8833 hCoV-19/England/QEUH-9F7B11/2020\|EPI_ISL_589236\|20 | ···················· | ···················· | ···················· | ···················· | ····T··············· | ···················· | |
| M | 8834 hCoV-19/England/MILK-9F2958/2020\|EPI_ISL_589237\|20 | ···················· | ···················· | ···················· | ···················· | ···················· | ···················· | |
| M | 8835 hCoV-19/England/QEUH-9F7D84/2020\|EPI_ISL_589238\|20 | ···················· | ···················· | ···················· | ···················· | ···················· | ···················· | |
| M | 8836 hCoV-19/England/QEUH-9CF550/2020\|EPI_ISL_590141\|20 | ···················· | ···················· | ···················· | ···················· | ···················· | ···················· | |
| M | 8837 hCoV-19/England/QEUH-9CE8B2/2020\|EPI_ISL_590142\|20 | ···················· | ···················· | ···················· | ···················· | ···················· | ···················· | |
| M | 8838 hCoV-19/England/QEUH-9CE849/2020\|EPI_ISL_590144\|20 | ···················· | ···················· | ···················· | ···················· | ···················· | ···················· | |
| M | 8839 hCoV-19/Wales/QEUH-9CE82B/2020\|EPI_ISL_590145\|2020 | ···················· | ···················· | ···················· | ···················· | ···················· | ···················· | |
| M | 8840 hCoV-19/England/QEUH-9CE83A/2020\|EPI_ISL_590146\|20 | ···················· | ···················· | ···················· | ···················· | ···················· | ···················· | |
| M | 8841 hCoV-19/England/QEUH-9CE1D5/2020\|EPI_ISL_590147\|20 | ···················· | ·A·················· | ···················· | ···················· | ···················· | ···················· | |
| M | 8842 hCoV-19/England/QEUH-9CE23C/2020\|EPI_ISL_590148\|20 | ···················· | ···················· | ···················· | ···················· | ···················· | ···················· | |
| M | 8843 hCoV-19/England/QEUH-9CF74B/2020\|EPI_ISL_590149\|20 | ···················· | ···················· | ···················· | ···················· | ···················· | ···················· | |
| M | 8844 hCoV-19/England/QEUH-9CF22C/2020\|EPI_ISL_590150\|20 | ···················· | ···················· | ···················· | ···················· | ···················· | ···················· | |
| M | 8845 hCoV-19/England/QEUH-9CE700/2020\|EPI_ISL_590151\|20 | ···················· | ···················· | ···················· | ···················· | ····T··············· | ···················· | |
| M | 8846 hCoV-19/England/ALDP-9B5C1C/2020\|EPI_ISL_590210\|20 | ···················· | ···················· | ···················· | ···················· | ···················· | ···················· | |
| M | 8847 hCoV-19/England/ALDP-9B58DC/2020\|EPI_ISL_590215\|20 | ···················· | ·A·················· | ···················· | ···················· | ···················· | ···A················ | |
| M | 8848 hCoV-19/England/QEUH-9B555D/2020\|EPI_ISL_590216\|20 | ···················· | ···················· | ···················· | ···················· | ···················· | ···················· | |
| M | 8849 hCoV-19/England/QEUH-9B4B10/2020\|EPI_ISL_590217\|20 | ···················· | ···················· | ···················· | ···················· | ···················· | ···················· | |
| M | 8850 hCoV-19/England/QEUH-9B4FD8/2020\|EPI_ISL_590218\|20 | ···················· | ···················· | ···················· | ···················· | ····T··············· | ···················· | |
| M | 8851 hCoV-19/England/QEUH-9B4961/2020\|EPI_ISL_590219\|20 | ···················· | ···················· | ···················· | ···················· | ···················· | ···················· | |
| M | 8852 hCoV-19/Japan/NF-02/2020\|EPI_ISL_591472\|2020-03 | ···················· | ···················· | ···················· | ···················· | ···················· | ···················· | |
| M | 8853 hCoV-19/Japan/NF-03/2020\|EPI_ISL_591473\|2020-03 | ···················· | ···················· | ···················· | ···················· | ···················· | ···················· | |
| M | 8854 hCoV-19/Japan/NF-04/2020\|EPI_ISL_591474\|2020-03 | ···················· | ···················· | ···················· | ···················· | ···················· | ···················· | |
| M | 8855 hCoV-19/Japan/NF-05/2020\|EPI_ISL_591475\|2020-03 | ···················· | ···················· | ···················· | ···················· | ···················· | ···················· | |
| M | 8856 hCoV-19/Japan/NF-06/2020\|EPI_ISL_591476\|2020-03 | ···················· | ···················· | ···················· | ···················· | ···················· | ···················· | |
| M | 8857 hCoV-19/Japan/NF-07/2020\|EPI_ISL_591477\|2020-03 | ···················· | ···················· | ···················· | ···················· | ···················· | ···················· | |
| M | 8858 hCoV-19/Japan/NF-08/2020\|EPI_ISL_591478\|2020-03 | ···················· | ···················· | ···················· | ···················· | ···················· | ···················· | |
| M | 8859 hCoV-19/Japan/NF-09/2020\|EPI_ISL_591479\|2020-04 | ···················· | ···················· | ···················· | ···················· | ···················· | ···················· | |
| M | 8860 hCoV-19/Japan/NF-10/2020\|EPI_ISL_591480\|2020-04 | ···················· | ···················· | ···················· | ···················· | ···················· | ···················· | |
| M | 8861 hCoV-19/Japan/NF-11/2020\|EPI_ISL_591481\|2020-04 | ···················· | ···················· | ···················· | ···················· | ···················· | ···················· | |
| M | 8862 hCoV-19/Japan/NF-12/2020\|EPI_ISL_591482\|2020-04 | ···················· | ···················· | ···················· | ···················· | ···················· | ···················· | |
| M | 8863 hCoV-19/France/OCC-33/2020\|EPI_ISL_591541\|2020-06- | ···················· | ···················· | ···················· | ···················· | ···················· | ···················· | |
| M | 8864 hCoV-19/France/OCC-38/2020\|EPI_ISL_591546\|2020-06- | ···················· | ···················· | ···················· | ···················· | ···················· | ···················· | |
| M | 8865 hCoV-19/France/OCC-39/2020\|EPI_ISL_591547\|2020-06- | ···················· | ···················· | ···················· | ···················· | ···················· | ···················· | |
| M | 8866 hCoV-19/Australia/VIC14514/2020\|EPI_ISL_592870\|202 | ···················· | ···················· | ···················· | ···················· | ···················· | ···················· | |
| M | 8867 hCoV-19/Australia/VIC14515/2020\|EPI_ISL_592871\|202 | ···················· | ···················· | ···················· | ···················· | ···················· | ···················· | |
| M | 8869 hCoV-19/Australia/VIC14525/2020\|EPI_ISL_592877\|202 | ···················· | ···················· | ···················· | ···················· | ···················· | ···················· | |
| M | 8870 hCoV-19/Australia/VIC14529/2020\|EPI_ISL_592878\|202 | ···················· | ···················· | ···················· | ···················· | ···················· | ···················· | |
| M | 8871 hCoV-19/Australia/VIC14531/2020\|EPI_ISL_592879\|202 | ···················· | ···················· | ···················· | ···················· | ···················· | ···················· | |
| M | 8873 hCoV-19/Australia/VIC14574/2020\|EPI_ISL_592901\|202 | ···················· | ···················· | ···················· | ···················· | ···················· | ···················· | |
| M | 8874 hCoV-19/Australia/VIC14575/2020\|EPI_ISL_592902\|202 | ···················· | ···················· | ···················· | ···················· | ···················· | ···················· | |
| M | 8875 hCoV-19/Australia/VIC14577/2020\|EPI_ISL_592903\|202 | ···················· | ···················· | ···················· | ···················· | ······R············· | ···················· | |
| M | 8876 hCoV-19/Australia/VIC14580/2020\|EPI_ISL_592904\|202 | ···················· | ···················· | ···················· | ···················· | ···················· | ···················· | |
| M | 8878 hCoV-19/Australia/VIC14589/2020\|EPI_ISL_592906\|202 | ···················· | ···················· | ···················· | ···················· | ···················· | ···················· | |
| M | 8882 hCoV-19/Australia/VIC14596/2020\|EPI_ISL_592910\|202 | ···················· | ···················· | ···················· | ···················· | ···················· | ···················· | |
| M | 8883 hCoV-19/Australia/VIC14597/2020\|EPI_ISL_592911\|202 | ···················· | ···················· | ···················· | ···················· | ···················· | ···················· | |
| M | 8884 hCoV-19/Australia/VIC14602/2020\|EPI_ISL_592912\|202 | ···················· | ···················· | ···················· | ···················· | ···················· | ···················· | |
| M | 8885 hCoV-19/Australia/VIC14607/2020\|EPI_ISL_592913\|202 | ···················· | ···················· | ···················· | ···················· | ···················· | ···················· | |
| M | 8887 hCoV-19/Australia/VIC14612/2020\|EPI_ISL_592915\|202 | ···················· | ···················· | ···················· | ···················· | ···················· | ···················· | |
| M | 8889 hCoV-19/Australia/VIC14615/2020\|EPI_ISL_592917\|202 | ···················· | ···················· | ···················· | ···················· | ···················· | ···················· | |
| M | 8891 hCoV-19/Australia/VIC14622/2020\|EPI_ISL_592919\|202 | ···················· | ···················· | ···················· | ···················· | ···················· | ···················· | |
| M | 8893 hCoV-19/Australia/VIC14625/2020\|EPI_ISL_592921\|202 | ···················· | ···················· | ···················· | ···················· | NNN················· | ···················· | 1 |
| M | 8895 hCoV-19/Australia/VIC14627/2020\|EPI_ISL_592923\|202 | ···················· | ···················· | ···················· | ···················· | ···················· | ···················· | |
| M | 8896 hCoV-19/Australia/VIC14628/2020\|EPI_ISL_592924\|202 | ···················· | ···················· | ···················· | ···················· | ···················· | ···················· | |
| M | 8897 hCoV-19/Australia/VIC14631/2020\|EPI_ISL_592925\|202 | ···················· | ···················· | ···················· | ···················· | ···················· | ···················· | |
| M | 8898 hCoV-19/Australia/VIC14633/2020\|EPI_ISL_592926\|202 | ···················· | ···················· | ···················· | ···················· | ···················· | ···················· | |
| M | 8899 hCoV-19/Australia/VIC14637/2020\|EPI_ISL_592927\|202 | ···················· | ···················· | ···················· | ···················· | ···················· | ···················· | |
| M | 8900 hCoV-19/Australia/VIC14638/2020\|EPI_ISL_592928\|202 | ···················· | ···················· | ···················· | ···················· | ···················· | ···················· | |
| M | 8902 hCoV-19/Australia/VIC14643/2020\|EPI_ISL_592930\|202 | ···················· | ···················· | ···················· | ···················· | ···················· | ···················· | |
| M | 8903 hCoV-19/Australia/VIC14646/2020\|EPI_ISL_592931\|202 | ···················· | ···················· | ···················· | ···················· | ···················· | ···················· | |
| M | 8904 hCoV-19/Australia/VIC14647/2020\|EPI_ISL_592932\|202 | ···················· | ···················· | ···················· | ···················· | ···················· | ···················· | |
| M | 8906 hCoV-19/Australia/VIC14650/2020\|EPI_ISL_592934\|202 | ···················· | ···················· | ···················· | ···················· | ···················· | ···················· | |
| M | 8907 hCoV-19/Australia/VIC14653/2020\|EPI_ISL_592935\|202 | ···················· | ···················· | ···················· | ···················· | ···················· | ···················· | |
| M | 8909 hCoV-19/Australia/VIC14657/2020\|EPI_ISL_592937\|202 | ···················· | ···················· | ···················· | ···················· | ···················· | ···················· | |
| M | 8910 hCoV-19/Australia/VIC14658/2020\|EPI_ISL_592938\|202 | ···················· | ···················· | ···················· | ···················· | ···················· | ···················· | |
| M | 8911 hCoV-19/Australia/VIC14660/2020\|EPI_ISL_592939\|202 | ···················· | ···················· | ···················· | ···················· | ···················· | ···················· | |
| M | 8912 hCoV-19/England/MILK-A23BDF/2020\|EPI_ISL_595105\|20 | ···················· | ···················· | ···················· | ···················· | ···················· | ···················· | |
| M | 8913 hCoV-19/England/MILK-A23BFD/2020\|EPI_ISL_595106\|20 | ···················· | ···················· | ···················· | ···················· | ···················· | ···················· | |
| M | 8914 hCoV-19/England/MILK-A23C36/2020\|EPI_ISL_595109\|20 | ···················· | ···················· | ···················· | ···················· | ···················· | ···················· | |
| M | 8915 hCoV-19/England/MILK-A23C45/2020\|EPI_ISL_595110\|20 | ···················· | ···················· | ···················· | ···················· | ···················· | ···················· | |

| | | | | | | | | |
|---|---|---|---|---|---|---|---|---|
| M | 8916 hCoV-19/England/MILK-A23C54/2020\|EPI_ISL_595111\|20 | ···················· | ···················· | ···················· | ···················· | ···················· | ···················· | |
| M | 8917 hCoV-19/England/MILK-A23C81/2020\|EPI_ISL_595114\|20 | ···················· | ···················· | ···················· | ···················· | ···················· | ···················· | |
| M | 8918 hCoV-19/England/MILK-A55E62/2020\|EPI_ISL_595115\|20 | ···················· | ···················· | ···················· | ···················· | ···················· | ···················· | |
| M | 8919 hCoV-19/England/MILK-A55E80/2020\|EPI_ISL_595116\|20 | ···················· | ·A·················· | ···················· | ···················· | ···················· | ···A················ | |
| M | 8920 hCoV-19/Wales/MILK-A55E9F/2020\|EPI_ISL_595117\|2020 | ···················· | ···················· | ···················· | ···················· | ···················· | ···················· | |
| M | 8921 hCoV-19/England/MILK-A55EDB/2020\|EPI_ISL_595118\|20 | ···················· | ···················· | ···················· | ···················· | ···················· | ···················· | |
| M | 8922 hCoV-19/England/MILK-A55F05/2020\|EPI_ISL_595119\|20 | ···················· | ···················· | ···················· | ···················· | ···················· | ···················· | |
| M | 8923 hCoV-19/England/MILK-A55F14/2020\|EPI_ISL_595120\|20 | ···················· | ···················· | ···················· | ···················· | ···················· | ···················· | |
| M | 8924 hCoV-19/England/MILK-A55F23/2020\|EPI_ISL_595121\|20 | ···················· | ···················· | ···················· | ···················· | NNN················· | ···················· | 1 |
| M | 8925 hCoV-19/England/MILK-A55F41/2020\|EPI_ISL_595122\|20 | ···················· | ···················· | ···················· | ···················· | ···················· | ···················· | |
| M | 8926 hCoV-19/England/MILK-A55FAB/2020\|EPI_ISL_595123\|20 | ···················· | ···················· | ···················· | ···················· | ···················· | ···················· | |
| M | 8927 hCoV-19/England/MILK-A55FBA/2020\|EPI_ISL_595124\|20 | ···················· | ···················· | ···················· | ···················· | ···················· | ···················· | |
| M | 8928 hCoV-19/Wales/MILK-A55FE7/2020\|EPI_ISL_595125\|2020 | ···················· | ···················· | ···················· | ···················· | ···················· | ···················· | |
| M | 8929 hCoV-19/England/MILK-A55FF6/2020\|EPI_ISL_595126\|20 | ···················· | ···················· | ···················· | ···················· | ······T············· | ···················· | |
| M | 8930 hCoV-19/England/MILK-A5602F/2020\|EPI_ISL_595127\|20 | ···················· | ···················· | ···················· | ···················· | ···················· | ···················· | |
| M | 8931 hCoV-19/England/MILK-A5604D/2020\|EPI_ISL_595129\|20 | ···················· | ···················· | ···················· | ···················· | ······T············· | ···················· | |
| M | 8933 hCoV-19/England/MILK-A5606B/2020\|EPI_ISL_595131\|20 | ···················· | ···················· | ···················· | ···················· | ······T············· | NNN················· | 1 |
| M | 8934 hCoV-19/England/MILK-A5607A/2020\|EPI_ISL_595132\|20 | ···················· | ···················· | ···················· | ···················· | ······T············· | ···················· | |
| M | 8935 hCoV-19/England/MILK-A56089/2020\|EPI_ISL_595133\|20 | ···················· | ·A·······A·········· | ···················· | ···················· | ···················· | ···················· | |
| M | 8936 hCoV-19/England/MILK-A560A7/2020\|EPI_ISL_595134\|20 | ···················· | ···················· | ···················· | ···················· | ···················· | ···················· | |
| M | 8937 hCoV-19/England/MILK-A5612C/2020\|EPI_ISL_595135\|20 | ···················· | ···················· | ···················· | ···················· | ······T············· | ···················· | |
| M | 8938 hCoV-19/England/MILK-A5621A/2020\|EPI_ISL_595140\|20 | ···················· | C··················· | ···················· | ···················· | ···················· | ···················· | |
| M | 8939 hCoV-19/Wales/MILK-A56238/2020\|EPI_ISL_595141\|2020 | ···················· | ···················· | ···················· | ···················· | ···················· | ···················· | |
| M | 8940 hCoV-19/Wales/MILK-A56247/2020\|EPI_ISL_595142\|2020 | ···················· | ···················· | ···················· | ···················· | ···················· | ···················· | |
| M | 8941 hCoV-19/Wales/MILK-A56256/2020\|EPI_ISL_595143\|2020 | ···················· | ···················· | ···················· | ···················· | ···················· | ···················· | |
| M | 8942 hCoV-19/Wales/MILK-A56274/2020\|EPI_ISL_595145\|2020 | ···················· | ···················· | ···················· | ···················· | ···················· | ···················· | |
| M | 8943 hCoV-19/Wales/MILK-A562A1/2020\|EPI_ISL_595146\|2020 | ···················· | ···················· | ···················· | ···················· | ···················· | ···················· | |
| M | 8944 hCoV-19/Wales/MILK-A562B0/2020\|EPI_ISL_595147\|2020 | ···················· | ···················· | ···················· | ···················· | ···················· | ···················· | |
| M | 8945 hCoV-19/Wales/MILK-A562DE/2020\|EPI_ISL_595148\|2020 | ···················· | ···················· | ···················· | ···················· | ···················· | ···················· | |
| M | 8946 hCoV-19/Wales/MILK-A562FC/2020\|EPI_ISL_595149\|2020 | ···················· | ···················· | ···················· | ···················· | ···················· | ···················· | |
| M | 8947 hCoV-19/Wales/MILK-A56371/2020\|EPI_ISL_595150\|2020 | ···················· | ···················· | ···················· | ···················· | ···················· | ···················· | |
| M | 8949 hCoV-19/Wales/MILK-A563BD/2020\|EPI_ISL_595152\|2020 | ···················· | ···················· | ···················· | ···················· | ···················· | ···················· | |
| M | 8950 hCoV-19/England/MILK-A56414/2020\|EPI_ISL_595155\|20 | ···················· | ···················· | ···················· | ···················· | ···················· | ···················· | |
| M | 8951 hCoV-19/England/MILK-A56441/2020\|EPI_ISL_595157\|20 | ···················· | ···················· | ···················· | ···················· | ···················· | ···················· | |
| M | 8954 hCoV-19/England/NORW-EFE2B/2020\|EPI_ISL_595162\|202 | ···················· | ···················· | ···················· | ···················· | ···················· | ···················· | |
| M | 8962 hCoV-19/England/ALDP-A2D7A4/2020\|EPI_ISL_586900\|20 | ···················· | ···················· | ···················· | ···················· | ···················· | ···················· | |
| M | 8963 hCoV-19/England/ALDP-A2DE81/2020\|EPI_ISL_586901\|20 | ···················· | ···················· | ···················· | ···················· | ···················· | ···················· | |
| M | 8964 hCoV-19/England/ALDP-A2E099/2020\|EPI_ISL_586903\|20 | ···················· | ···················· | ···················· | ···················· | ···················· | ···················· | |
| M | 8965 hCoV-19/England/ALDP-A2D8DE/2020\|EPI_ISL_586904\|20 | ···················· | ···················· | ···················· | ···················· | ···················· | ···················· | |
| M | 8966 hCoV-19/England/ALDP-A2DA9C/2020\|EPI_ISL_586905\|20 | ···················· | ···················· | ···················· | ···················· | ···················· | ···················· | |
| M | 8967 hCoV-19/England/ALDP-A2E4F7/2020\|EPI_ISL_586906\|20 | ···················· | ···················· | ···················· | ···················· | ···················· | ···················· | |
| M | 8968 hCoV-19/England/ALDP-A2E05D/2020\|EPI_ISL_586908\|20 | ···················· | ···················· | ···················· | ···················· | ···················· | ···················· | |
| M | 8969 hCoV-19/England/ALDP-A2DD75/2020\|EPI_ISL_586909\|20 | ···················· | ···················· | ···················· | ···················· | ···················· | ···················· | |
| M | 8970 hCoV-19/England/ALDP-A2D0C7/2020\|EPI_ISL_586910\|20 | ···················· | ···················· | ···················· | ···················· | ···················· | ···················· | |
| M | 8971 hCoV-19/England/ALDP-A2DE90/2020\|EPI_ISL_586911\|20 | ···················· | ···················· | ···················· | ···················· | ···················· | ···················· | |
| M | 8972 hCoV-19/England/ALDP-A2D838/2020\|EPI_ISL_586912\|20 | ···················· | ···················· | ···················· | ···················· | ···················· | ···················· | |
| M | 8973 hCoV-19/England/ALDP-A2E70D/2020\|EPI_ISL_586913\|20 | ···················· | ···················· | ···················· | ···················· | ···················· | ···················· | |
| M | 8974 hCoV-19/England/ALDP-A2D3FB/2020\|EPI_ISL_586914\|20 | ···················· | ···················· | ···················· | ···················· | ···················· | ···················· | |
| M | 8975 hCoV-19/England/ALDP-A2D71D/2020\|EPI_ISL_586915\|20 | ···················· | ···················· | ···················· | ···················· | ···················· | ···················· | |
| M | 8976 hCoV-19/England/ALDP-A2D7FF/2020\|EPI_ISL_586916\|20 | ···················· | ···················· | ···················· | ···················· | ···················· | ···················· | |
| M | 8977 hCoV-19/England/ALDP-A2E15A/2020\|EPI_ISL_586917\|20 | ···················· | ···················· | ···················· | ···················· | ···················· | ···················· | |
| M | 8978 hCoV-19/England/ALDP-A2D364/2020\|EPI_ISL_586918\|20 | ···················· | ···················· | ···················· | ···················· | ···················· | ···················· | |
| M | 8979 hCoV-19/England/ALDP-A2E22A/2020\|EPI_ISL_586919\|20 | ···················· | ···················· | ···················· | ···················· | ···················· | ···················· | |
| M | 8980 hCoV-19/England/ALDP-A2DFAC/2020\|EPI_ISL_586920\|20 | ···················· | ···················· | ···················· | ···················· | ···················· | ···················· | |
| M | 8981 hCoV-19/England/ALDP-A2E5F4/2020\|EPI_ISL_586921\|20 | ···················· | ···················· | ···················· | ···················· | ···················· | ···················· | |
| M | 8982 hCoV-19/England/ALDP-A2E460/2020\|EPI_ISL_586922\|20 | ···················· | ·A·················· | ···················· | ···················· | ···················· | ···················· | |
| M | 8983 hCoV-19/England/ALDP-A2E64C/2020\|EPI_ISL_586923\|20 | ···················· | ···················· | ···················· | ···················· | ···················· | ···················· | |
| M | 8984 hCoV-19/England/ALDP-A2E5E5/2020\|EPI_ISL_586924\|20 | ···················· | ···················· | ···················· | ···················· | ···················· | ···A················ | |
| M | 8985 hCoV-19/England/ALDP-A2DB8A/2020\|EPI_ISL_586925\|20 | ···················· | ···················· | ···················· | ···················· | ···················· | ···················· | |
| M | 8986 hCoV-19/Wales/ALDP-A2D513/2020\|EPI_ISL_586926\|2020 | ···················· | ···················· | ···················· | ···················· | ···················· | ···················· | |
| M | 8987 hCoV-19/England/ALDP-A2DC2D/2020\|EPI_ISL_586927\|20 | ···················· | ···················· | ···················· | ···················· | ···················· | ···················· | |
| M | 8988 hCoV-19/England/ALDP-A2D81A/2020\|EPI_ISL_586928\|20 | ···················· | ···················· | ···················· | ···················· | ···················· | ···················· | |
| M | 8989 hCoV-19/England/ALDP-A2E293/2020\|EPI_ISL_586929\|20 | ···················· | ···················· | ···················· | ···················· | ···················· | ···················· | |
| M | 8990 hCoV-19/England/ALDP-A2DAC9/2020\|EPI_ISL_586930\|20 | ···················· | ···················· | ···················· | ···················· | ···················· | ···················· | |
| M | 8991 hCoV-19/England/ALDP-A2E65B/2020\|EPI_ISL_586931\|20 | ···················· | ···················· | ···················· | ···················· | ···················· | ···················· | |
| M | 8992 hCoV-19/England/ALDP-A2DB02/2020\|EPI_ISL_586932\|20 | ···················· | ···················· | ···················· | ···················· | ···················· | ···················· | |
| M | 8993 hCoV-19/England/ALDP-A2DE09/2020\|EPI_ISL_586933\|20 | ···················· | ···················· | ···················· | ···················· | ···················· | ···················· | |
| M | 8994 hCoV-19/England/ALDP-A2D795/2020\|EPI_ISL_586934\|20 | ···················· | ···················· | ···················· | ···················· | ···················· | ···················· | |
| M | 8995 hCoV-19/England/ALDP-A2D601/2020\|EPI_ISL_586935\|20 | ···················· | ···················· | ···················· | ···················· | ···················· | ···················· | |
| M | 8996 hCoV-19/England/ALDP-A2E7EF/2020\|EPI_ISL_586936\|20 | ···················· | ···················· | ···················· | ···················· | ···················· | ···················· | |
| M | 8997 hCoV-19/England/ALDP-A2E20C/2020\|EPI_ISL_586937\|20 | ···················· | ···················· | ···················· | ···················· | ···················· | ···················· | |
| M | 8998 hCoV-19/England/ALDP-A2FB00/2020\|EPI_ISL_586938\|20 | ···················· | ···················· | ···················· | ···················· | ···················· | ···················· | |
| M | 8999 hCoV-19/England/ALDP-A2EC1D/2020\|EPI_ISL_586939\|20 | ···················· | ···················· | ···················· | ···················· | ···················· | ···················· | |
| M | 9000 hCoV-19/England/MILK-9F274F/2020\|EPI_ISL_589101\|20 | ···················· | ···················· | ···················· | ···················· | ···················· | ···················· | |
| M | 9001 hCoV-19/Northern Ireland/QEUH-9F9915/2020\|EPI_ISL_ | ···················· | ···················· | ···················· | ···················· | ···················· | ···················· | |
| M | 9002 hCoV-19/England/MILK-9F242A/2020\|EPI_ISL_589118\|20 | ···················· | ···················· | ···················· | ···················· | ···················· | ···················· | |
| M | 9003 hCoV-19/England/QEUH-9F97DF/2020\|EPI_ISL_589119\|20 | ···················· | ···················· | ···················· | ···················· | ···················· | ···················· | |
| M | 9004 hCoV-19/England/QEUH-9F9809/2020\|EPI_ISL_589120\|20 | ···················· | ···················· | ···················· | ···················· | ···················· | ···················· | |
| M | 9005 hCoV-19/England/MILK-9F28D3/2020\|EPI_ISL_589121\|20 | ···················· | ···················· | ···················· | ···················· | ···················· | ···················· | |
| M | 9006 hCoV-19/England/MILK-9F2A37/2020\|EPI_ISL_589123\|20 | ···················· | ·A·······A·········· | ···················· | ···················· | ···················· | ···················· | |

M 9007 hCoV-19/England/QEUH-9F74CB/2020|EPI_ISL_589124|20 ···················· ···················· ···················· ···················· ···················· ····················
M 9008 hCoV-19/England/QEUH-9F9C1C/2020|EPI_ISL_589125|20 ···················· ·A·················· ···················· ···················· ···················· ··A·················
M 9009 hCoV-19/England/QEUH-9F9784/2020|EPI_ISL_589126|20 ···················· ···················· ···················· ···················· ···················· ····················
M 9010 hCoV-19/England/QEUH-9F74DA/2020|EPI_ISL_589128|20 ···················· ···················· ···················· ···················· ···················· ····················
M 9011 hCoV-19/England/QEUH-9F7C1E/2020|EPI_ISL_589129|20 ···················· ···················· ···················· ···················· ···T················ ····················
M 9012 hCoV-19/England/QEUH-9F7B5D/2020|EPI_ISL_589131|20 ···················· ···················· ···················· ···················· ···················· ····················
M 9013 hCoV-19/England/QEUH-9F962D/2020|EPI_ISL_589132|20 ···················· ···················· ···················· ···················· ···················· ····················
M 9014 hCoV-19/England/MILK-9F29C1/2020|EPI_ISL_589133|20 ···················· ·A·················· ···················· ···················· ···················· ··A·················
M 9015 hCoV-19/England/QEUH-9F7E45/2020|EPI_ISL_589136|20 ···················· ···················· ···················· ···················· ···················· ····················
M 9016 hCoV-19/England/QEUH-9F78ED/2020|EPI_ISL_589138|20 ···················· ···················· ···················· ···················· ···················· ····················
M 9017 hCoV-19/England/QEUH-9F7434/2020|EPI_ISL_589139|20 ···················· ···················· ···················· ···················· ···················· ····················
M 9018 hCoV-19/England/QEUH-9F749E/2020|EPI_ISL_589140|20 ···················· ···················· ···················· ···················· ···················· ····················
M 9019 hCoV-19/England/QEUH-9F98FA/2020|EPI_ISL_589141|20 ···················· ···················· ···················· ···················· ···················· ····················
M 9020 hCoV-19/England/QEUH-9F95E4/2020|EPI_ISL_589142|20 ···················· ···················· ···················· ···················· ···················· ····················
M 9021 hCoV-19/England/QEUH-9F7B99/2020|EPI_ISL_589144|20 ···················· ···················· ···················· ···················· ···T················ ····················
M 9022 hCoV-19/England/QEUH-9F957B/2020|EPI_ISL_589145|20 ···················· ···················· ···················· ···················· ···················· ····················
M 9023 hCoV-19/England/QEUH-9F99CA/2020|EPI_ISL_589146|20 ···················· ···················· ···················· ···················· ···················· ····················
M 9024 hCoV-19/England/MILK-9F2527/2020|EPI_ISL_589147|20 ···················· ···················· ···················· ···················· ···················· ····················
M 9025 hCoV-19/England/QEUH-9F7847/2020|EPI_ISL_589148|20 ···················· ···················· ···················· ···················· ···T················ ····················
M 9026 hCoV-19/England/QEUH-9F74E9/2020|EPI_ISL_589150|20 ···················· ···················· ···················· ···················· ···················· ····················
M 9027 hCoV-19/England/QEUH-9F9827/2020|EPI_ISL_589151|20 ···················· ···················· ···················· ···················· ···················· ····················
M 9028 hCoV-19/England/QEUH-9F9687/2020|EPI_ISL_589152|20 ···················· ···················· ···················· ···················· ···················· ····················
M 9029 hCoV-19/England/MILK-9F20BD/2020|EPI_ISL_589153|20 ···················· ···················· ···················· ···················· ···················· ····················
M 9030 hCoV-19/England/QEUH-9F7E09/2020|EPI_ISL_589154|20 ···················· ·A·················· ···················· ···················· ···················· ····················
M 9031 hCoV-19/England/QEUH-9F78A1/2020|EPI_ISL_589155|20 ···················· ···················· ···················· ···················· ···················· ····················
M 9032 hCoV-19/England/QEUH-9F963C/2020|EPI_ISL_589156|20 ···················· ···················· ···················· ···················· ···················· ····················
M 9033 hCoV-19/England/MILK-9F2AFB/2020|EPI_ISL_589157|20 ···················· ···················· ···················· ···················· ···················· ····················
M 9034 hCoV-19/England/QEUH-9F7601/2020|EPI_ISL_589158|20 ···················· ···················· ···················· ···················· ···················· ····················
M 9035 hCoV-19/England/MILK-9F216F/2020|EPI_ISL_589170|20 ···················· ···················· ···················· ···················· ···················· ····················
M 9036 hCoV-19/England/MILK-9F2105/2020|EPI_ISL_589171|20 ···················· ···················· ···················· ···················· ···················· ····················
M 9037 hCoV-19/England/MILK-9F22B7/2020|EPI_ISL_589172|20 ···················· ···················· ···················· ···················· ···················· ····················
M 9038 hCoV-19/England/MILK-9F22D5/2020|EPI_ISL_589173|20 ···················· ···················· ···················· ···················· ···················· ····················
M 9039 hCoV-19/Wales/MILK-9F26E8/2020|EPI_ISL_589174|2020 ···················· ···················· ···················· ···················· ···················· ····················
M 9040 hCoV-19/England/QEUH-9F7452/2020|EPI_ISL_589175|20 ···················· ···················· ···················· ···················· ···················· ····················
M 9041 hCoV-19/England/MILK-9F2211/2020|EPI_ISL_589176|20 ···················· ···················· ···················· ···················· ···················· ····················
M 9042 hCoV-19/England/QEUH-9F9906/2020|EPI_ISL_589177|20 ···················· ···················· ···················· ···················· ···················· ····················
M 9043 hCoV-19/England/QEUH-9F77FF/2020|EPI_ISL_589178|20 ···················· ···················· ···················· ···················· ···T················ ····················
M 9044 hCoV-19/Wales/MILK-9F2A82/2020|EPI_ISL_589179|2020 ···················· ···················· ···················· ···················· ···················· ····················
M 9045 hCoV-19/England/QEUH-9F7522/2020|EPI_ISL_589180|20 ···················· ···················· ···················· ···················· ···················· ····················
M 9046 hCoV-19/England/QEUH-9F7A50/2020|EPI_ISL_589181|20 ···················· ···················· ···················· ···················· ···················· ····················
M 9047 hCoV-19/England/MILK-9F2044/2020|EPI_ISL_589183|20 ···················· ···················· ···················· ···················· ···················· ····················
M 9048 hCoV-19/England/MILK-9F2B43/2020|EPI_ISL_589185|20 ···················· ···················· ···················· ···················· ···················· ····················
M 9049 hCoV-19/England/QEUH-9F7C3C/2020|EPI_ISL_589186|20 ···················· ···················· ···················· ···················· ···················· ····················
M 9050 hCoV-19/England/QEUH-9F954E/2020|EPI_ISL_589188|20 ···················· ···················· ···················· ···················· ···················· ····················
M 9051 hCoV-19/England/QEUH-9F7E36/2020|EPI_ISL_589189|20 ···················· ···················· ···················· ···················· ···················· ····················
M 9052 hCoV-19/England/QEUH-9F9CC1/2020|EPI_ISL_589190|20 ···················· ···················· ···················· ···················· ···················· ····················
M 9053 hCoV-19/England/QEUH-9F96E1/2020|EPI_ISL_589191|20 ···················· ···················· ···················· ···················· ·····T·············· ····················
M 9054 hCoV-19/England/QEUH-9F9A6D/2020|EPI_ISL_589192|20 ···················· ·A·················· ···················· ···················· ···················· ····················
M 9055 hCoV-19/England/QEUH-9F7AC9/2020|EPI_ISL_589194|20 ···················· ···················· ···················· ···················· ···················· ····················
M 9056 hCoV-19/England/MILK-9F276D/2020|EPI_ISL_589195|20 ···················· ·A·················· ···················· ···················· ···················· ··A·················
M 9057 hCoV-19/Wales/MILK-9F2A28/2020|EPI_ISL_589196|2020 ···················· ···················· ···················· ···················· ···················· ····················
M 9058 hCoV-19/England/QEUH-9F95A8/2020|EPI_ISL_589197|20 ···················· ···················· ···················· ···················· ···················· ····················
M 9059 hCoV-19/England/MILK-9F22A8/2020|EPI_ISL_589199|20 ···················· ···················· ···················· ···················· ···················· ····················
M 9060 hCoV-19/England/QEUH-9CF417/2020|EPI_ISL_590100|20 ···················· ···················· ···················· ···················· ···················· ····················
M 9061 hCoV-19/England/QEUH-9CE21E/2020|EPI_ISL_590101|20 ···················· ···················· ···················· ···················· ···················· ····················
M 9062 hCoV-19/England/QEUH-9CF7C3/2020|EPI_ISL_590102|20 ···················· ···················· ···················· ···················· ···················· ····················
M 9063 hCoV-19/England/QEUH-9CE7B5/2020|EPI_ISL_590103|20 ···················· ···················· ···················· ···················· ···················· ····················
M 9064 hCoV-19/England/QEUH-9CE858/2020|EPI_ISL_590104|20 ···················· ···················· ···················· ···················· ···················· ····················
M 9065 hCoV-19/England/QEUH-9CEB22/2020|EPI_ISL_590105|20 ···················· ···················· ···················· ···················· ···················· ····················
M 9066 hCoV-19/England/QEUH-9CF839/2020|EPI_ISL_590107|20 ···················· ···················· ···················· ···················· ···················· ····················
M 9067 hCoV-19/England/QEUH-9CE560/2020|EPI_ISL_590108|20 ···················· ···················· ···················· ···················· ···················· ····················
M 9068 hCoV-19/England/QEUH-9CED77/2020|EPI_ISL_590109|20 ···················· ·A·················· ···················· ···················· ···················· ····················
M 9069 hCoV-19/England/QEUH-9CF56F/2020|EPI_ISL_590110|20 ···················· ···················· ···················· ···················· ···················· ····················
M 9070 hCoV-19/England/QEUH-9CF73C/2020|EPI_ISL_590111|20 ···················· ···················· ···················· ···················· ···················· ····················
M 9071 hCoV-19/England/QEUH-9CF15C/2020|EPI_ISL_590112|20 ···················· ···················· ···················· ···················· ···················· ····················
M 9072 hCoV-19/England/QEUH-9CE1B7/2020|EPI_ISL_590113|20 ···················· ·A·················· ···················· ···················· ···················· ····················
M 9073 hCoV-19/England/QEUH-9CE490/2020|EPI_ISL_590114|20 ···················· ···················· ···················· ···················· ···················· ····················
M 9074 hCoV-19/England/QEUH-9CE375/2020|EPI_ISL_590115|20 ···················· ···················· ···················· ···················· ···················· ····················
M 9075 hCoV-19/England/QEUH-9CE58E/2020|EPI_ISL_590116|20 ···················· ···················· ···················· ···················· ···················· ····················
M 9076 hCoV-19/England/QEUH-9CEDE0/2020|EPI_ISL_590117|20 ···················· ·A·················· ···················· ···················· ···················· ··A·················
M 9077 hCoV-19/England/QEUH-9CEBF5/2020|EPI_ISL_590118|20 ···················· ···················· ···················· ···················· ···················· ····················
M 9078 hCoV-19/England/QEUH-9CECF2/2020|EPI_ISL_590120|20 ···················· ···················· ···················· ···················· ···················· ····················
M 9079 hCoV-19/England/QEUH-9CF0AA/2020|EPI_ISL_590121|20 ···················· ····A··············· ···················· ···················· ···················· ····················
M 9080 hCoV-19/England/QEUH-9CF295/2020|EPI_ISL_590122|20 ···················· ···················· ···················· ···················· ···················· ····················
M 9081 hCoV-19/England/QEUH-9CED2C/2020|EPI_ISL_590123|20 ···················· ···················· ···················· ···················· ···················· ····················
M 9082 hCoV-19/England/QEUH-9CE9FB/2020|EPI_ISL_590124|20 ···················· ···················· ···················· ···················· ···················· ····················
M 9083 hCoV-19/England/QEUH-9CF408/2020|EPI_ISL_590125|20 ···················· ···················· ···················· ···················· ···················· ····················
M 9084 hCoV-19/England/QEUH-9CEC10/2020|EPI_ISL_590127|20 ···················· ·A·················· ···················· ···················· ···················· ····················
M 9085 hCoV-19/England/QEUH-9CE22D/2020|EPI_ISL_590128|20 ···················· ···················· ···················· ···················· ···T················ ····················
M 9086 hCoV-19/England/QEUH-9CED1D/2020|EPI_ISL_590129|20 ···················· ···················· ···················· ···················· ···················· ····················

M 9087 hCoV-19/England/QEUH-9CE919/2020|EPI_ISL_590130|20 ···················· ·A·················· ···················· ···················· ···················· ····················
M 9088 hCoV-19/England/QEUH-9CF981/2020|EPI_ISL_590131|20 ···················· ···················· ···················· ···················· ···················· ····················
M 9089 hCoV-19/England/QEUH-9CE6D6/2020|EPI_ISL_590132|20 ···················· ···················· ···················· ···················· ···················· ····················
M 9090 hCoV-19/England/QEUH-9CF462/2020|EPI_ISL_590133|20 ···················· ···················· ···················· ···················· ······T············· ····················
M 9091 hCoV-19/England/QEUH-9CEE74/2020|EPI_ISL_590134|20 ···················· ···················· ···················· ···················· ···················· ····················
M 9092 hCoV-19/England/QEUH-9CF9BE/2020|EPI_ISL_590135|20 ···················· ···················· ···················· ···················· ···················· ····················
M 9093 hCoV-19/England/QEUH-9CED0E/2020|EPI_ISL_590136|20 ···················· ···················· ···················· ···················· ····T··············· ····················
M 9094 hCoV-19/England/QEUH-9CF259/2020|EPI_ISL_590137|20 ···················· ···················· ···················· ···················· ···················· ····················
M 9095 hCoV-19/England/QEUH-9CE4CD/2020|EPI_ISL_590138|20 ···················· ···················· ···················· ···················· ···················· ····················
M 9096 hCoV-19/England/QEUH-9CEC3E/2020|EPI_ISL_590139|20 ···················· ·A·················· ···················· ···················· ···················· ····················
M 9097 hCoV-19/England/QEUH-9CE436/2020|EPI_ISL_590140|20 ···················· ···················· ···················· ···················· ···················· ····················
M 9098 hCoV-19/England/QEUH-9CF435/2020|EPI_ISL_590152|20 ···················· ···················· ···················· ···················· ···················· ····················
M 9099 hCoV-19/England/QEUH-9CE8A3/2020|EPI_ISL_590153|20 ···················· ···················· ···················· ···················· ···················· ····················
M 9100 hCoV-19/England/QEUH-9CED68/2020|EPI_ISL_590154|20 ···················· ···················· ···················· ···················· ····T··············· ····················
M 9101 hCoV-19/England/QEUH-9CE876/2020|EPI_ISL_590155|20 ···················· ···················· ···················· ···················· ···················· ····················
M 9102 hCoV-19/England/QEUH-9CE72E/2020|EPI_ISL_590156|20 ···················· ···················· ···················· ···················· ···················· ····················
M 9103 hCoV-19/England/QEUH-9CF2A4/2020|EPI_ISL_590157|20 ···················· ···················· ···················· ···················· ···················· ····················
M 9104 hCoV-19/England/QEUH-9CF71E/2020|EPI_ISL_590158|20 ···················· ···················· ···················· ···················· ···················· ····················
M 9105 hCoV-19/England/QEUH-9CF5D8/2020|EPI_ISL_590159|20 ···················· ···················· ···················· ···················· ···················· ····················
M 9106 hCoV-19/England/QEUH-9CE5CA/2020|EPI_ISL_590160|20 ···················· ···················· ···················· ···················· ···················· ····················
M 9107 hCoV-19/England/QEUH-9CF5E7/2020|EPI_ISL_590161|20 ···················· ···················· ···················· ···················· ···················· ····················
M 9108 hCoV-19/England/QEUH-9CE32A/2020|EPI_ISL_590162|20 ···················· ···················· ···················· ···················· ···················· ····················
M 9109 hCoV-19/England/QEUH-9CE4AF/2020|EPI_ISL_590163|20 ···················· ·A·················· ···················· ···················· ···················· ····················
M 9110 hCoV-19/England/QEUH-9CF927/2020|EPI_ISL_590164|20 ···················· ···················· ···················· ···················· ···················· ····················
M 9111 hCoV-19/England/QEUH-9CF75A/2020|EPI_ISL_590165|20 ···················· ···················· ···················· ···················· ···················· ····················
M 9112 hCoV-19/England/QEUH-9CE8C1/2020|EPI_ISL_590166|20 ···················· ···················· ···················· ···················· ····T··············· ····················
M 9113 hCoV-19/England/QEUH-9CF031/2020|EPI_ISL_590168|20 ···················· ···················· ···················· ···················· ···················· ····················
M 9114 hCoV-19/England/QEUH-9CE76A/2020|EPI_ISL_590169|20 ···················· ·A·················· ···················· ···················· ···················· ···A················
M 9115 hCoV-19/England/QEUH-9CE65E/2020|EPI_ISL_590170|20 ···················· ···················· ···················· ···················· ····T··············· ····················
M 9116 hCoV-19/England/QEUH-9CEB9B/2020|EPI_ISL_590171|20 ···················· ···················· ···················· ···················· ···················· ····················
M 9117 hCoV-19/Wales/MILK-9C2F04/2020|EPI_ISL_590172|2020 ···················· ···················· ···················· ···················· ···················· ····················
M 9118 hCoV-19/Wales/CAMC-9B1AAD/2020|EPI_ISL_590173|2020 ···················· ···················· ···················· ···················· ···················· ····················
M 9119 hCoV-19/Wales/MILK-9C2414/2020|EPI_ISL_590174|2020 ···················· ···················· ···················· ···················· ···················· ····················
M 9120 hCoV-19/Wales/MILK-9C3B2D/2020|EPI_ISL_590175|2020 ···················· ···················· ···················· ···················· ···················· ····················
M 9121 hCoV-19/Wales/QEUH-9C7B56/2020|EPI_ISL_590176|2020 ···················· ···················· ···················· ···················· ···················· ····················
M 9122 hCoV-19/Wales/QEUH-9C98F3/2020|EPI_ISL_590177|2020 ···················· ···················· ···················· ···················· ···················· ····················
M 9123 hCoV-19/England/QEUH-9B4E26/2020|EPI_ISL_590178|20 ···················· ···················· ···················· ···················· ···················· ····················
M 9124 hCoV-19/England/QEUH-9B4B89/2020|EPI_ISL_590179|20 ···················· ···················· ···················· ···················· ···················· ····················
M 9125 hCoV-19/England/QEUH-9B4D92/2020|EPI_ISL_590180|20 ···················· ···················· ···················· ···················· ···················· ····················
M 9126 hCoV-19/England/ALDP-9B5836/2020|EPI_ISL_590181|20 ···················· ···················· ···················· ···················· ···················· ····················
M 9127 hCoV-19/England/QEUH-9B5001/2020|EPI_ISL_590182|20 ···················· ···················· ···················· ···················· ···················· ····················
M 9128 hCoV-19/England/QEUH-9B5098/2020|EPI_ISL_590183|20 ···················· ···················· ···················· ···················· ···················· ····················
M 9129 hCoV-19/England/QEUH-9B54AB/2020|EPI_ISL_590184|20 ···················· ···················· ···················· ···················· ···················· ····················
M 9130 hCoV-19/England/QEUH-9B4A8C/2020|EPI_ISL_590185|20 ···················· ···················· ···················· ···················· ···················· ····················
M 9131 hCoV-19/England/ALDP-9B5AE5/2020|EPI_ISL_590186|20 ···················· ···················· ···················· ···················· ···················· ····················
M 9132 hCoV-19/England/QEUH-9B56D2/2020|EPI_ISL_590187|20 ···················· ·A·················· ···················· ···················· ···················· ···A················
M 9133 hCoV-19/England/QEUH-9B4A22/2020|EPI_ISL_590188|20 ···················· ···················· ···················· ···················· ···················· ····················
M 9134 hCoV-19/England/QEUH-9B466A/2020|EPI_ISL_590189|20 ···················· ···················· ···················· ···················· ···················· ····················
M 9135 hCoV-19/England/QEUH-9B4A31/2020|EPI_ISL_590190|20 ···················· ···················· ···················· ···················· ···················· ····················
M 9136 hCoV-19/England/QEUH-9B4EBD/2020|EPI_ISL_590191|20 ···················· ···················· ···················· ···················· ···················· ····················
M 9137 hCoV-19/England/QEUH-9B512C/2020|EPI_ISL_590192|20 ···················· ···················· ···················· ···················· ···················· ····················
M 9138 hCoV-19/England/ALDP-9B5C3A/2020|EPI_ISL_590193|20 ···················· ···················· ···················· ···················· ···················· ····················
M 9139 hCoV-19/England/ALDP-9B5890/2020|EPI_ISL_590194|20 ···················· ···················· ···················· ···················· ···················· ····················
M 9140 hCoV-19/England/QEUH-9B51E0/2020|EPI_ISL_590195|20 ···················· ···················· ···················· ···················· ···················· ····················
M 9141 hCoV-19/England/QEUH-9B46F1/2020|EPI_ISL_590196|20 ···················· ···················· ···················· ···················· ···················· ····················
M 9142 hCoV-19/England/QEUH-9B507A/2020|EPI_ISL_590197|20 ···················· ···················· ···················· ···················· ···················· ····················
M 9143 hCoV-19/England/ALDP-9B58EB/2020|EPI_ISL_590198|20 ···················· ···················· ···················· ···················· ···················· ····················
M 9144 hCoV-19/England/QEUH-9B4688/2020|EPI_ISL_590199|20 ···················· ···················· ···················· ···················· ···················· ····················
M 9145 hCoV-19/Japan/IC-0058/2020|EPI_ISL_591400|2020-06 ···················· ···················· ···················· ···················· ···················· ····················
M 9146 hCoV-19/Japan/IC-0059/2020|EPI_ISL_591401|2020-06 ···················· ···················· ···················· ···················· ···················· ····················
M 9147 hCoV-19/Japan/IC-0060/2020|EPI_ISL_591402|2020-06 ···················· ···················· ···················· ···················· ···················· ····················
M 9148 hCoV-19/Japan/IC-0061/2020|EPI_ISL_591403|2020-06 ···················· ···················· ···················· ···················· ···················· ····················
M 9149 hCoV-19/Japan/IC-0062/2020|EPI_ISL_591404|2020-06 ···················· ·A·················· ···················· ···················· ···················· ···A················
M 9150 hCoV-19/Japan/IC-0063/2020|EPI_ISL_591405|2020-06 ···················· ···················· ···················· ···················· ···················· ····················
M 9151 hCoV-19/Japan/IC-0064/2020|EPI_ISL_591406|2020-06 ···················· ····A··············· ···················· ···················· ···················· ····················
M 9152 hCoV-19/Japan/IC-0065/2020|EPI_ISL_591407|2020-06 ···················· ·A·················· ···················· ···················· ···················· ···A················
M 9153 hCoV-19/Japan/IC-0066/2020|EPI_ISL_591408|2020-06 ···················· ···················· ···················· ···················· ···················· ····················
M 9154 hCoV-19/Japan/IC-0067/2020|EPI_ISL_591409|2020-06 ···················· ···················· ···················· ···················· ···················· ····················
M 9155 hCoV-19/Japan/IC-0068/2020|EPI_ISL_591410|2020-06 ···················· ···················· ···················· ···················· ···················· ····················
M 9156 hCoV-19/Japan/IC-0069/2020|EPI_ISL_591411|2020-07 ···················· ···················· ···················· ···················· ···················· ····················
M 9157 hCoV-19/Japan/IC-0071/2020|EPI_ISL_591413|2020-07 ···················· ·A·················· ···················· ···················· ···················· ····················
M 9158 hCoV-19/Japan/IC-0088/2020|EPI_ISL_591430|2020-07 ···················· ······A············· ···················· ···················· ···················· ····················
M 9159 hCoV-19/Japan/IC-0089/2020|EPI_ISL_591431|2020-07 ···················· ···················· ···················· ···················· ···················· ····················
M 9160 hCoV-19/Japan/IC-0091/2020|EPI_ISL_591432|2020-07 ···················· ···················· ···················· ···················· ···················· ····················
M 9161 hCoV-19/Japan/IC-0093/2020|EPI_ISL_591434|2020-07 ···················· ···················· ···················· ···················· ···················· ····················
M 9162 hCoV-19/Japan/IC-0094/2020|EPI_ISL_591435|2020-07 ···················· ···················· ···················· ···················· ···················· ····················
M 9163 hCoV-19/Japan/IC-0095/2020|EPI_ISL_591436|2020-07 ···················· ···················· ···················· ···················· ···················· ····················
M 9164 hCoV-19/Japan/IC-0096/2020|EPI_ISL_591437|2020-07 ···················· ···················· ···················· ···················· ···················· ····················
M 9165 hCoV-19/Japan/IC-0097/2020|EPI_ISL_591438|2020-07 ···················· ···················· ···················· ···················· ···················· ····················
M 9166 hCoV-19/Japan/IC-0098/2020|EPI_ISL_591439|2020-07 ···················· ···················· ···················· ···················· ···················· ····················

```
M 9167 hCoV-19/Japan/IC-0099/2020|EPI_ISL_591440|2020-07 ···················· ···················· ····················· ···················· ···················· ····················
M 9168 hCoV-19/Japan/IC-0100/2020|EPI_ISL_591441|2020-07 ···················· ···················· ····················· ···················· ···················· ····················
M 9169 hCoV-19/Japan/IC-0101/2020|EPI_ISL_591442|2020-07 ···················· ···················· ····················· ···················· ···················· ····················
M 9170 hCoV-19/Japan/IC-0102/2020|EPI_ISL_591443|2020-07 ···················· ···················· ····················· ···················· ···················· ····················
M 9171 hCoV-19/Japan/IC-0103/2020|EPI_ISL_591444|2020-07 ···················· ·A·················· ····················· ···················· ···················· ··A·················
M 9172 hCoV-19/Japan/IC-0104/2020|EPI_ISL_591445|2020-07 ···················· ···················· ····················· ···················· ···················· ····················
M 9173 hCoV-19/Japan/IC-0105/2020|EPI_ISL_591446|2020-07 ···················· ···················· ····················· ···················· ···················· ····················
M 9174 hCoV-19/Japan/IC-0106/2020|EPI_ISL_591447|2020-07 ···················· ···················· ····················· ···················· ···················· ····················
M 9175 hCoV-19/Japan/IC-0107/2020|EPI_ISL_591448|2020-07 ···················· ···················· ····················· ···················· ···················· ····················
M 9176 hCoV-19/Japan/IC-0108/2020|EPI_ISL_591449|2020-07 ···················· ···················· ····················· ···················· ···················· ····················
M 9177 hCoV-19/Japan/IC-0109/2020|EPI_ISL_591450|2020-07 ···················· ···················· ····················· ···················· ···················· ····················
M 9178 hCoV-19/Japan/IC-0110/2020|EPI_ISL_591451|2020-07 ···················· ···················· ····················· ···················· ···················· ····················
M 9179 hCoV-19/Japan/IC-0111/2020|EPI_ISL_591452|2020-07 ···················· ···················· ····················· ···················· ···················· ····················
M 9180 hCoV-19/Japan/IC-0112/2020|EPI_ISL_591453|2020-07 ···················· ···················· ····················· ···················· ···················· ····················
M 9181 hCoV-19/Japan/IC-0113/2020|EPI_ISL_591454|2020-07 ···················· ···················· ····················· ···················· ···················· ····················
M 9182 hCoV-19/Japan/IC-0114/2020|EPI_ISL_591455|2020-07 ···················· ···················· ····················· ···················· ···················· ····················
M 9183 hCoV-19/Japan/IC-0115/2020|EPI_ISL_591456|2020-07 ···················· ···················· ····················· ···················· ···················· ····················
M 9184 hCoV-19/Japan/IC-0116/2020|EPI_ISL_591457|2020-07 ···················· ···················· ····················· ···················· ···················· ····················
M 9185 hCoV-19/Japan/IC-0117/2020|EPI_ISL_591458|2020-07 ···················· ···················· ····················· ···················· ···················· ····················
M 9186 hCoV-19/Japan/IC-0118/2020|EPI_ISL_591459|2020-07 ···················· ···················· ····················· ···················· ···················· ····················
M 9187 hCoV-19/Japan/IC-0119/2020|EPI_ISL_591460|2020-08 ···················· ···················· ····················· ···················· ···················· ····················
M 9188 hCoV-19/Japan/IC-0120/2020|EPI_ISL_591461|2020-08 ···················· ···················· ····················· ···················· ···················· ····················
M 9189 hCoV-19/Japan/IC-0121/2020|EPI_ISL_591462|2020-08 ···················· ···················· ····················· ···················· ···················· ····················
M 9190 hCoV-19/Japan/IC-0122/2020|EPI_ISL_591463|2020-08 ···················· ···················· ····················· ···················· ···················· ····················
M 9191 hCoV-19/Japan/IC-0123/2020|EPI_ISL_591464|2020-08 ···················· ···················· ····················· ···················· ···················· ····················
M 9192 hCoV-19/Japan/IC-0124/2020|EPI_ISL_591465|2020-08 ···················· ···················· ····················· ···················· ···················· ····················
M 9193 hCoV-19/Japan/IC-0125/2020|EPI_ISL_591466|2020-08 ···················· ···················· ····················· ···················· ···················· ····················
M 9194 hCoV-19/Japan/IC-0126/2020|EPI_ISL_591467|2020-08 ···················· ···················· ····················· ···················· ···················· ····················
M 9195 hCoV-19/Japan/IC-0127/2020|EPI_ISL_591468|2020-08 ···················· ···················· ····················· ···················· ···················· ····················
M 9196 hCoV-19/Japan/IC-0128/2020|EPI_ISL_591469|2020-08 ···················· ···················· ····················· ···················· ···················· ····················
M 9197 hCoV-19/Japan/IC-0129/2020|EPI_ISL_591470|2020-08 ···················· ···················· ····················· ···················· ···················· ····················
M 9198 hCoV-19/Japan/NF-01/2020|EPI_ISL_591471|2020-03 ···················· ···················· ····················· ···················· ···················· ····················
M 9199 hCoV-19/Japan/NF-13/2020|EPI_ISL_591483|2020-04 ···················· ···················· ····················· ···················· ···················· ····················
M 9200 hCoV-19/Australia/NSW985/2020|EPI_ISL_591484|2020- ···················· ···················· ····················· ···················· ···················· ····················
M 9201 hCoV-19/Australia/NSW1012/2020|EPI_ISL_591485|2020 ···················· ···················· ····················· ···················· ···················· ····················
M 9202 hCoV-19/Australia/NSW1014/2020|EPI_ISL_591486|2020 ···················· ···················· ····················· ···················· ···················· ····················
M 9203 hCoV-19/Australia/NSW987/2020|EPI_ISL_591487|2020- ···················· ···················· ····················· ···················· ···················· ····················
M 9204 hCoV-19/Australia/NSW998/2020|EPI_ISL_591488|2020- ···················· ···················· ····················· ···················· ···················· ····················
M 9205 hCoV-19/Australia/NSW1009/2020|EPI_ISL_591489|2020 ···················· ···················· ····················· ···················· ···················· ····················
M 9206 hCoV-19/Australia/NSW1010/2020|EPI_ISL_591490|2020 ···················· ···················· ····················· ···················· ···················· ····················
M 9207 hCoV-19/Australia/NSW1011/2020|EPI_ISL_591491|2020 ···················· ···················· ····················· ···················· ···················· ····················
M 9208 hCoV-19/Australia/NSW1003/2020|EPI_ISL_591492|2020 ···················· ···················· ····················· ···················· ···················· ····················
M 9209 hCoV-19/Australia/NSW988/2020|EPI_ISL_591493|2020- ···················· ···················· ····················· ···················· ···················· ····················
M 9210 hCoV-19/Australia/NSW981/2020|EPI_ISL_591494|2020- ···················· ···················· ····················· ···················· ···················· ····················
M 9211 hCoV-19/Australia/NSW982/2020|EPI_ISL_591495|2020- ···················· ···················· ····················· ···················· ···················· ····················
M 9212 hCoV-19/Australia/NSW991/2020|EPI_ISL_591496|2020- ········T··········· ···················· ····················· ···················· ···················· ····················
M 9213 hCoV-19/Australia/NSW995/2020|EPI_ISL_591497|2020- ···················· ···················· ····················· ···················· ···················· ····················
M 9214 hCoV-19/Australia/NSW996/2020|EPI_ISL_591498|2020- ···················· ···················· ····················· ···················· ···················· ····················
M 9215 hCoV-19/Australia/NSW997/2020|EPI_ISL_591499|2020- ···················· ···················· ····················· ···················· ···················· ····················
M 9216 hCoV-19/Australia/VIC14552/2020|EPI_ISL_592704|202 ···················· ···················· ····················· ···················· ···················· ····················
M 9217 hCoV-19/England/CAMB-1B6712/2020|EPI_ISL_584300|20 ···················· ···················· ····················· ···················· ···················· ····················
M 9218 hCoV-19/England/CAMB-1B676D/2020|EPI_ISL_584303|20 ···················· ···················· ····················· ···················· ···················· ····················
M 9223 hCoV-19/England/CAMB-1B68A6/2020|EPI_ISL_584311|20 ···················· ···················· ····················· ···················· ···················· ····················
M 9224 hCoV-19/England/CAMB-1B68B5/2020|EPI_ISL_584312|20 ···················· ···················· ····················· ···················· ···················· ····················
M 9225 hCoV-19/England/CAMB-1B68C4/2020|EPI_ISL_584313|20 ···················· ···················· ····················· ···················· ···················· ····················
M 9228 hCoV-19/England/CAMB-1B691C/2020|EPI_ISL_584316|20 ···················· ···················· ····················· ···················· ···················· ····················
M 9230 hCoV-19/England/CAMB-1B6976/2020|EPI_ISL_584319|20 ···················· ···················· ····················· ···················· ···················· ····················
M 9233 hCoV-19/England/CAMC-9D70AF/2020|EPI_ISL_584344|20 ···················· ···················· ····················· ···················· ···················· ····················
M 9234 hCoV-19/England/CAMC-9D70BE/2020|EPI_ISL_584345|20 ···················· ·A·················· ····················· ···················· ···················· ··A·················
M 9235 hCoV-19/England/CAMC-9D70FA/2020|EPI_ISL_584346|20 ···················· ···················· ····················· ···················· ···················· ····················
M 9236 hCoV-19/England/CAMC-9D7133/2020|EPI_ISL_584347|20 ···················· ···················· ····················· ···················· ···················· ····················
M 9237 hCoV-19/England/CAMC-9D7160/2020|EPI_ISL_584349|20 ···················· ···················· ····················· ···················· ···················· ····················
M 9238 hCoV-19/England/CAMC-9D719D/2020|EPI_ISL_584350|20 ···················· ···················· ····················· ···················· ···················· ····················
M 9239 hCoV-19/England/CAMC-9D73C4/2020|EPI_ISL_584366|20 ···················· ···················· ····················· ···················· ···················· ····················
M 9240 hCoV-19/England/CAMC-9D73D3/2020|EPI_ISL_584367|20 ···················· ···················· ····················· ···················· ···················· ····················
M 9241 hCoV-19/England/CAMC-9D73F1/2020|EPI_ISL_584368|20 ···················· ···················· ····················· ···················· ···················· ····················
M 9242 hCoV-19/England/CAMC-9D741C/2020|EPI_ISL_584369|20 ···················· ···················· ····················· ···················· ······T············· ····················
M 9243 hCoV-19/England/CAMC-9D743A/2020|EPI_ISL_584370|20 ···················· ···················· ····················· ···················· ···················· ····················
M 9244 hCoV-19/England/CAMC-9D7458/2020|EPI_ISL_584371|20 ···················· ···················· ····················· ···················· ···················· ····················
M 9245 hCoV-19/England/CAMC-9D7467/2020|EPI_ISL_584372|20 ···················· ···················· ····················· ···················· ···················· ····················
M 9246 hCoV-19/England/CAMC-9D7476/2020|EPI_ISL_584373|20 ···················· ·A·················· ····················· ···················· ···················· ··A·················
M 9247 hCoV-19/England/CAMC-9D74B2/2020|EPI_ISL_584375|20 ···················· ·A·················· ····················· ···················· ···················· ··A·················
M 9248 hCoV-19/England/CAMC-9D74FE/2020|EPI_ISL_584377|20 ···················· ···················· ····················· ···················· ···················· ····················
M 9249 hCoV-19/England/CAMC-9D7537/2020|EPI_ISL_584378|20 ···················· ··---··············· ····················· ···················· ···················· ····················
M 9250 hCoV-19/England/CAMC-9D7564/2020|EPI_ISL_584379|20 ···················· ···················· ····················· ···················· ···················· ····················
M 9251 hCoV-19/England/CAMC-9D7582/2020|EPI_ISL_584380|20 ···················· ···················· ····················· ···················· ···················· ····················
M 9252 hCoV-19/England/CAMC-9EB8CE/2020|EPI_ISL_584382|20 ···················· ·A·················· ····················· ···················· ···················· ··A·················
M 9253 hCoV-19/England/CAMC-9EB916/2020|EPI_ISL_584384|20 ···················· ·A·················· ····················· ···················· ···················· ····················
M 9254 hCoV-19/England/CAMC-9EB9CB/2020|EPI_ISL_584386|20 ···················· ···················· ····················· ···················· ···················· ····················
M 9255 hCoV-19/England/CAMC-9EB9E9/2020|EPI_ISL_584387|20 ···················· ···················· ····················· ···················· ···················· ····················
```

| | | | | | | | | | |
|---|---|---|---|---|---|---|---|---|---|
| M | 9256 | hCoV-19/England/CAMC-9EBA22/2020\|EPI_ISL_584388\|20 | ···················· | ···················· | ···················· | ···················· | ···················· | ···················· | |
| M | 9257 | hCoV-19/England/CAMC-9EBA31/2020\|EPI_ISL_584389\|20 | ···················· | ···················· | ···················· | ···················· | ···················· | ···················· | |
| M | 9258 | hCoV-19/England/CAMC-9EBA40/2020\|EPI_ISL_584390\|20 | ···················· | ···················· | ···················· | ···················· | ···················· | ···················· | |
| M | 9259 | hCoV-19/England/CAMC-9EBA5F/2020\|EPI_ISL_584391\|20 | ···················· | ···················· | ···················· | ···················· | ···················· | ···················· | |
| M | 9260 | hCoV-19/England/CAMC-9EBA6E/2020\|EPI_ISL_584392\|20 | ···················· | ···················· | ···················· | ···················· | ···················· | ···················· | |
| M | 9261 | hCoV-19/England/CAMC-9EBA7D/2020\|EPI_ISL_584393\|20 | ···················· | ·A·················· | ···················· | ···················· | ···················· | ··A················· | |
| M | 9262 | hCoV-19/England/CAMC-9EBA8C/2020\|EPI_ISL_584394\|20 | ···················· | ···················· | ···················· | ···················· | ···················· | ···················· | |
| M | 9263 | hCoV-19/England/CAMC-9EBAAA/2020\|EPI_ISL_584396\|20 | ···················· | ···················· | ···················· | ···················· | ···················· | ···················· | |
| M | 9264 | hCoV-19/England/CAMC-9EBB2F/2020\|EPI_ISL_584398\|20 | ···················· | ···················· | ···················· | ···················· | ···················· | ···················· | |
| M | 9266 | hCoV-19/Wales/CAMC-9D6DF4/2020\|EPI_ISL_585620\|2020 | ···················· | ···················· | ···················· | ···················· | ···················· | ···················· | |
| M | 9267 | hCoV-19/Wales/CAMC-9D6E1F/2020\|EPI_ISL_585621\|2020 | ···················· | ···················· | ···················· | ···················· | ···················· | ···················· | |
| M | 9268 | hCoV-19/Wales/CAMC-9D6F2B/2020\|EPI_ISL_585622\|2020 | ···················· | ···················· | ···················· | ···················· | ···················· | ···················· | |
| M | 9269 | hCoV-19/Wales/CAMC-9D7009/2020\|EPI_ISL_585623\|2020 | ···················· | ···················· | ···················· | ···················· | ···················· | ···················· | |
| M | 9270 | hCoV-19/Wales/CAMC-9D7018/2020\|EPI_ISL_585624\|2020 | ···················· | ···················· | ···················· | ···················· | ···················· | ···················· | |
| M | 9271 | hCoV-19/Wales/CAMC-9D70EB/2020\|EPI_ISL_585625\|2020 | ···················· | ···················· | ···················· | ···················· | ···················· | ···················· | |
| M | 9272 | hCoV-19/Wales/CAMC-9EBEAE/2020\|EPI_ISL_585627\|2020 | ···················· | ···················· | ···················· | ···················· | ···················· | ···················· | |
| M | 9273 | hCoV-19/Wales/CAMC-9EBF9C/2020\|EPI_ISL_585628\|2020 | ···················· | ···················· | ···················· | ···················· | ···················· | ···················· | |
| M | 9274 | hCoV-19/Wales/PHWC-16DE51/2020\|EPI_ISL_585630\|2020 | ···················· | ···················· | ···················· | ···················· | ···················· | ···················· | |
| M | 9276 | hCoV-19/Wales/PHWC-16DE7F/2020\|EPI_ISL_585632\|2020 | ···················· | ···················· | ···················· | ···················· | ···················· | ···················· | |
| M | 9277 | hCoV-19/Wales/PHWC-16DE8E/2020\|EPI_ISL_585633\|2020 | ···················· | ···················· | ···················· | ···················· | ···················· | ···················· | |
| M | 9279 | hCoV-19/Wales/PHWC-16DEAC/2020\|EPI_ISL_585635\|2020 | ···················· | ···················· | ···················· | ···················· | ···················· | ···················· | |
| M | 9280 | hCoV-19/Wales/PHWC-16DEBB/2020\|EPI_ISL_585636\|2020 | ···················· | ···················· | ···················· | ···················· | ···················· | ···················· | |
| M | 9285 | hCoV-19/Wales/PHWC-16DF6D/2020\|EPI_ISL_585645\|2020 | ···················· | ···················· | ···················· | ···················· | ···················· | ···················· | |
| M | 9287 | hCoV-19/Wales/PHWC-16DFB8/2020\|EPI_ISL_585649\|2020 | ···················· | ···················· | ···················· | ···················· | ···················· | ···················· | |
| M | 9288 | hCoV-19/Wales/PHWC-16DFC7/2020\|EPI_ISL_585650\|2020 | ···················· | ···················· | ···················· | ···················· | ···················· | ···················· | |
| M | 9292 | hCoV-19/Wales/PHWC-16E184/2020\|EPI_ISL_585677\|2020 | ···················· | ···················· | ···················· | ···················· | NNN················· | ···················· | 1 |
| M | 9293 | hCoV-19/Wales/PHWC-16E193/2020\|EPI_ISL_585678\|2020 | ···················· | ···················· | ···················· | ···················· | ···················· | ···················· | |
| M | 9297 | hCoV-19/Wales/PHWC-16E2EB/2020\|EPI_ISL_585698\|2020 | ···················· | ···················· | ···················· | ···················· | ···················· | ···················· | |
| M | 9298 | hCoV-19/Wales/PHWC-16E2FA/2020\|EPI_ISL_585699\|2020 | ···················· | ···················· | ···················· | ···················· | ···················· | ···················· | |
| M | 9299 | hCoV-19/England/ALDP-A2FD64/2020\|EPI_ISL_586940\|20 | ···················· | ···················· | ···················· | ···················· | ···················· | ···················· | |
| M | 9300 | hCoV-19/England/ALDP-A2E970/2020\|EPI_ISL_586941\|20 | ···················· | ·A·······A·········· | ···················· | ···················· | ···················· | ···················· | |
| M | 9301 | hCoV-19/England/ALDP-A2EA9B/2020\|EPI_ISL_586942\|20 | ···················· | ···················· | ···················· | ···················· | ···················· | ···················· | |
| M | 9302 | hCoV-19/England/ALDP-A2EE80/2020\|EPI_ISL_586943\|20 | ···················· | ···················· | ···················· | ···················· | ···················· | ···················· | |
| M | 9303 | hCoV-19/England/ALDP-A2F56C/2020\|EPI_ISL_586944\|20 | ···················· | ···················· | ···················· | ···················· | ···················· | ···················· | |
| M | 9304 | hCoV-19/England/ALDP-A2F1E0/2020\|EPI_ISL_586945\|20 | ···················· | ·A·················· | ···················· | ···················· | ···················· | ··A················· | |
| M | 9305 | hCoV-19/England/ALDP-A2F2DE/2020\|EPI_ISL_586946\|20 | ···················· | ···················· | ···················· | ···················· | ···················· | ···················· | |
| M | 9306 | hCoV-19/England/ALDP-A2FC1C/2020\|EPI_ISL_586947\|20 | ···················· | ···················· | ···················· | ···················· | ···················· | ···················· | |
| M | 9307 | hCoV-19/England/ALDP-A2E925/2020\|EPI_ISL_586948\|20 | ···················· | ···················· | ···················· | ···················· | ···················· | ···················· | |
| M | 9308 | hCoV-19/England/ALDP-A2FD46/2020\|EPI_ISL_586949\|20 | ···················· | ···················· | ···················· | ···················· | ···················· | ···················· | |
| M | 9309 | hCoV-19/England/ALDP-A2E9BC/2020\|EPI_ISL_586950\|20 | ···················· | ···················· | ···················· | ···················· | ···················· | ···················· | |
| M | 9310 | hCoV-19/England/ALDP-A2F317/2020\|EPI_ISL_586951\|20 | ···················· | ···················· | ···················· | ···················· | ···················· | ···················· | |
| M | 9311 | hCoV-19/England/ALDP-A2FA7C/2020\|EPI_ISL_586952\|20 | ···················· | ···················· | ···················· | ···················· | ···················· | ···················· | |
| M | 9312 | hCoV-19/England/ALDP-A2EE35/2020\|EPI_ISL_586953\|20 | ···················· | ·A·················· | ···················· | ···················· | ···················· | ···················· | |
| M | 9313 | hCoV-19/England/ALDP-A2F53F/2020\|EPI_ISL_586954\|20 | ···················· | ···················· | ···················· | ···················· | ···················· | ···················· | |
| M | 9314 | hCoV-19/England/ALDP-A2FB1F/2020\|EPI_ISL_586955\|20 | ···················· | ···················· | ···················· | ···················· | ···················· | ···················· | |
| M | 9315 | hCoV-19/England/ALDP-A2EA13/2020\|EPI_ISL_586956\|20 | ···················· | ···················· | ···················· | ···················· | ···················· | ···················· | |
| M | 9316 | hCoV-19/England/ALDP-A2F4D8/2020\|EPI_ISL_586957\|20 | ···················· | ···················· | ···················· | ···················· | ······T············· | ···················· | |
| M | 9317 | hCoV-19/England/ALDP-A2F195/2020\|EPI_ISL_586958\|20 | ···················· | ···················· | ···················· | ···················· | ···················· | ···················· | |
| M | 9318 | hCoV-19/England/ALDP-A2ED38/2020\|EPI_ISL_586959\|20 | ···················· | ···················· | ···················· | ···················· | ···················· | ···················· | |
| M | 9319 | hCoV-19/England/ALDP-A2EFAB/2020\|EPI_ISL_586960\|20 | ···················· | ···················· | ···················· | ···················· | ···················· | ···················· | |
| M | 9320 | hCoV-19/England/ALDP-A2EA5F/2020\|EPI_ISL_586961\|20 | ···················· | ···················· | ···················· | ···················· | ···················· | ···················· | |
| M | 9321 | hCoV-19/England/ALDP-A2E8CE/2020\|EPI_ISL_586962\|20 | ···················· | ·A·················· | ···················· | ···················· | ···················· | ··A················· | |
| M | 9322 | hCoV-19/England/ALDP-A2F793/2020\|EPI_ISL_586963\|20 | ···················· | ···················· | ···················· | ···················· | ···················· | ···················· | |
| M | 9323 | hCoV-19/England/ALDP-A2E907/2020\|EPI_ISL_586964\|20 | ···················· | ···················· | ···················· | ···················· | ···················· | ···················· | |
| M | 9324 | hCoV-19/England/ALDP-A2F924/2020\|EPI_ISL_586965\|20 | ···················· | ·A·················· | ···················· | ···················· | ···················· | ··A················· | |
| M | 9325 | hCoV-19/England/ALDP-A2EF41/2020\|EPI_ISL_586966\|20 | ···················· | ···················· | ···················· | ···················· | ···················· | ···················· | |
| M | 9326 | hCoV-19/England/ALDP-A2FE52/2020\|EPI_ISL_586967\|20 | ···················· | ···················· | ···················· | ···················· | ···················· | ···················· | |
| M | 9327 | hCoV-19/England/ALDP-A2FA4F/2020\|EPI_ISL_586968\|20 | ···················· | ···················· | ···················· | ···················· | ···················· | ···················· | |
| M | 9328 | hCoV-19/England/ALDP-A2E9DA/2020\|EPI_ISL_586969\|20 | ···················· | ···················· | ···················· | ···················· | ···················· | ···················· | |
| M | 9329 | hCoV-19/England/ALDP-A2EA22/2020\|EPI_ISL_586970\|20 | ···················· | ···················· | ···················· | ···················· | ···················· | ···················· | |
| M | 9330 | hCoV-19/England/ALDP-A2FBE2/2020\|EPI_ISL_586971\|20 | ···················· | ···················· | ···················· | ···················· | ···················· | ···················· | |
| M | 9331 | hCoV-19/England/ALDP-A2ED47/2020\|EPI_ISL_586972\|20 | ···················· | ·A·················· | ···················· | ···················· | ···················· | ··A················· | |
| M | 9332 | hCoV-19/England/ALDP-A2E6B5/2020\|EPI_ISL_586973\|20 | ···················· | ···················· | ···················· | ···················· | ···················· | ···················· | |
| M | 9333 | hCoV-19/England/ALDP-A2FBF1/2020\|EPI_ISL_586974\|20 | ···················· | ·A·················· | ···················· | ···················· | ···················· | ···················· | |
| M | 9334 | hCoV-19/England/ALDP-A2F6F0/2020\|EPI_ISL_586975\|20 | ···················· | ···················· | ···················· | ···················· | ···················· | ···················· | |
| M | 9335 | hCoV-19/England/ALDP-A2EDCF/2020\|EPI_ISL_586976\|20 | ···················· | ···················· | ···················· | ···················· | ···················· | ···················· | |
| M | 9336 | hCoV-19/England/ALDP-A2EEBD/2020\|EPI_ISL_586977\|20 | ···················· | ···················· | ···················· | ···················· | ···················· | ···················· | |
| M | 9337 | hCoV-19/England/ALDP-A2EBF2/2020\|EPI_ISL_586978\|20 | ···················· | ···················· | ···················· | ···················· | ···················· | ···················· | |
| M | 9338 | hCoV-19/England/ALDP-A2EAE6/2020\|EPI_ISL_586979\|20 | ···················· | ···················· | ···················· | ···················· | ···················· | ···················· | |
| M | 9339 | hCoV-19/England/ALDP-A2E9E9/2020\|EPI_ISL_586980\|20 | ···················· | ···················· | ···················· | ···················· | ···················· | ···················· | |
| M | 9340 | hCoV-19/England/ALDP-A2F9CA/2020\|EPI_ISL_586981\|20 | ···················· | ···················· | ···················· | ···················· | ···················· | ···················· | |
| M | 9341 | hCoV-19/England/ALDP-A2EB89/2020\|EPI_ISL_586982\|20 | ···················· | ·A·················· | ···················· | ···················· | ···················· | ··A················· | |
| M | 9342 | hCoV-19/England/ALDP-A2F186/2020\|EPI_ISL_586983\|20 | ···················· | ···················· | ···················· | ···················· | ···················· | ···················· | |
| M | 9343 | hCoV-19/England/QEUH-9F7ABA/2020\|EPI_ISL_589100\|20 | ···················· | ···················· | ···················· | ···················· | ···················· | ···················· | |
| M | 9344 | hCoV-19/England/MILK-9F2071/2020\|EPI_ISL_589103\|20 | ···················· | ···················· | ···················· | ···················· | ···················· | ···················· | |
| M | 9345 | hCoV-19/England/QEUH-9F7DDF/2020\|EPI_ISL_589105\|20 | ···················· | ···················· | ···················· | ···················· | ···················· | ···················· | |
| M | 9346 | hCoV-19/England/QEUH-9F7C4B/2020\|EPI_ISL_589106\|20 | ···················· | ···················· | ···················· | ···················· | ···················· | ···················· | |
| M | 9347 | hCoV-19/England/QEUH-9F7A9C/2020\|EPI_ISL_589107\|20 | ···················· | ···················· | ···················· | ···················· | ···················· | ···················· | |
| M | 9348 | hCoV-19/England/MILK-9F226C/2020\|EPI_ISL_589110\|20 | ···················· | ···················· | ···················· | ···················· | ···················· | ···················· | |
| M | 9349 | hCoV-19/England/MILK-9F2026/2020\|EPI_ISL_589111\|20 | ···················· | ···················· | ···················· | ···················· | ···················· | ···················· | |

| | | | | | | | |
|---|---|---|---|---|---|---|---|
| M | 9350 hCoV-19/England/QEUH-9F7E63/2020\|EPI_ISL_589112\|20 | ···················· | ···················· | ····················· | ···················· | ···················· | ···················· |
| M | 9351 hCoV-19/England/QEUH-9F716A/2020\|EPI_ISL_589113\|20 | ···················· | ···················· | ····················· | ···················· | ···················· | ···················· |
| M | 9352 hCoV-19/England/MILK-9F224E/2020\|EPI_ISL_589114\|20 | ···················· | ···················· | ····················· | ···················· | ···················· | ···················· |
| M | 9353 hCoV-19/England/MILK-9F1FC0/2020\|EPI_ISL_589115\|20 | ···················· | ···················· | ····················· | ···················· | ···················· | ···················· |
| M | 9354 hCoV-19/England/MILK-9F28F1/2020\|EPI_ISL_589117\|20 | ···················· | ···················· | ····················· | ···················· | ···················· | ···················· |
| M | 9355 hCoV-19/Japan/IC-0070/2020\|EPI_ISL_591412\|2020-07 | ···················· | ·A·················· | ····················· | ···················· | ···················· | ···················· |
| M | 9356 hCoV-19/Japan/IC-0072/2020\|EPI_ISL_591414\|2020-07 | ···················· | ···················· | ····················· | ···················· | ···················· | ···················· |
| M | 9357 hCoV-19/Japan/IC-0073/2020\|EPI_ISL_591415\|2020-07 | ···················· | ···················· | ····················· | ···················· | ···················· | ···················· |
| M | 9358 hCoV-19/Japan/IC-0074/2020\|EPI_ISL_591416\|2020-07 | ···················· | ···················· | ····················· | ···················· | ···················· | ···················· |
| M | 9359 hCoV-19/Japan/IC-0075/2020\|EPI_ISL_591417\|2020-07 | ···················· | ···················· | ····················· | ···················· | ···················· | ···················· |
| M | 9360 hCoV-19/Japan/IC-0076/2020\|EPI_ISL_591418\|2020-07 | ···················· | ···················· | ····················· | ···················· | ···················· | ···················· |
| M | 9361 hCoV-19/Japan/IC-0077/2020\|EPI_ISL_591419\|2020-07 | ···················· | ···················· | ····················· | ···················· | ···················· | ···················· |
| M | 9362 hCoV-19/Japan/IC-0078/2020\|EPI_ISL_591420\|2020-07 | ···················· | ···················· | ····················· | ···················· | ···················· | ···················· |
| M | 9363 hCoV-19/Japan/IC-0079/2020\|EPI_ISL_591421\|2020-07 | ···················· | ···················· | ····················· | ···················· | ···················· | ···················· |
| M | 9364 hCoV-19/Japan/IC-0080/2020\|EPI_ISL_591422\|2020-07 | ···················· | ···················· | ····················· | ···················· | ···················· | ···················· |
| M | 9365 hCoV-19/Japan/IC-0081/2020\|EPI_ISL_591423\|2020-07 | ···················· | ···················· | ····················· | ···················· | ···················· | ···················· |
| M | 9366 hCoV-19/Japan/IC-0082/2020\|EPI_ISL_591424\|2020-07 | ···················· | ···················· | ····················· | ···················· | ···················· | ···················· |
| M | 9367 hCoV-19/Japan/IC-0083/2020\|EPI_ISL_591425\|2020-07 | ···················· | ···················· | ····················· | ···················· | ···················· | ···················· |
| M | 9368 hCoV-19/Japan/IC-0084/2020\|EPI_ISL_591426\|2020-07 | ···················· | ···················· | ····················· | ···················· | ···················· | ···················· |
| M | 9369 hCoV-19/Japan/IC-0085/2020\|EPI_ISL_591427\|2020-07 | ···················· | ···················· | ····················· | ···················· | ···················· | ···················· |
| M | 9370 hCoV-19/Japan/IC-0086/2020\|EPI_ISL_591428\|2020-07 | ···················· | ···················· | ····················· | ···················· | ······T············· | ···················· |
| M | 9371 hCoV-19/Japan/IC-0087/2020\|EPI_ISL_591429\|2020-07 | ···················· | ···················· | ····················· | ···················· | ···················· | ···················· |
| M | 9373 hCoV-19/England/MILK-A17F2D/2020\|EPI_ISL_595000\|20 | ···················· | ···················· | ····················· | ···················· | ···················· | ···················· |
| M | 9374 hCoV-19/England/MILK-A17F78/2020\|EPI_ISL_595002\|20 | ···················· | ···················· | ····················· | ···················· | ······T············· | ···················· |
| M | 9375 hCoV-19/Wales/MILK-A18065/2020\|EPI_ISL_595014\|2020 | ···················· | ···················· | ····················· | ···················· | ···················· | ···················· |
| M | 9376 hCoV-19/England/MILK-A180CF/2020\|EPI_ISL_595016\|20 | ···················· | ···················· | ····················· | ···················· | ···················· | ···················· |
| M | 9377 hCoV-19/England/MILK-A180DE/2020\|EPI_ISL_595017\|20 | ···················· | ···················· | ····················· | ···················· | ···················· | ···················· |
| M | 9378 hCoV-19/England/MILK-A18162/2020\|EPI_ISL_595021\|20 | ···················· | ···················· | ····················· | ···················· | ···················· | ···················· |
| M | 9379 hCoV-19/England/MILK-A1829C/2020\|EPI_ISL_595023\|20 | ···················· | ···················· | ····················· | ···················· | ···················· | ···················· |
| M | 9380 hCoV-19/England/MILK-A1836C/2020\|EPI_ISL_595025\|20 | ···················· | ···················· | ····················· | ···················· | ···················· | ···················· |
| M | 9381 hCoV-19/England/MILK-A183C6/2020\|EPI_ISL_595031\|20 | ···················· | ···················· | ····················· | ···················· | ···················· | ···················· |
| M | 9382 hCoV-19/England/MILK-A1843C/2020\|EPI_ISL_595033\|20 | ···················· | ···················· | ····················· | ···················· | ···················· | ···················· |
| M | 9383 hCoV-19/England/MILK-A18469/2020\|EPI_ISL_595035\|20 | ···················· | ···················· | ····················· | ···················· | ···················· | ···················· |
| M | 9384 hCoV-19/England/MILK-A18487/2020\|EPI_ISL_595037\|20 | ···················· | ···················· | ····················· | ···················· | ···················· | ···················· |
| M | 9385 hCoV-19/England/MILK-A18496/2020\|EPI_ISL_595038\|20 | ···················· | ···················· | ····················· | ···················· | ···················· | ···················· |
| M | 9386 hCoV-19/England/MILK-A184D2/2020\|EPI_ISL_595041\|20 | ···················· | ···················· | ····················· | ···················· | ···················· | ···················· |
| M | 9387 hCoV-19/England/MILK-A184E1/2020\|EPI_ISL_595042\|20 | ···················· | ···················· | ····················· | ···················· | ···················· | ···················· |
| M | 9388 hCoV-19/England/MILK-A184F0/2020\|EPI_ISL_595043\|20 | ···················· | ···················· | ····················· | ···················· | ···················· | ···················· |
| M | 9389 hCoV-19/England/MILK-A1851B/2020\|EPI_ISL_595044\|20 | ···················· | ···················· | ····················· | ···················· | ···················· | ···················· |
| M | 9390 hCoV-19/England/MILK-A1852A/2020\|EPI_ISL_595045\|20 | ···················· | ·A·················· | ····················· | ···················· | ···················· | ···················· |
| M | 9391 hCoV-19/England/MILK-A1D2D3/2020\|EPI_ISL_595046\|20 | ···················· | ···················· | ····················· | ···················· | ···················· | ···················· |
| M | 9392 hCoV-19/England/MILK-A1D2E2/2020\|EPI_ISL_595047\|20 | ···················· | ···················· | ····················· | ···················· | ···················· | ···················· |
| M | 9393 hCoV-19/England/MILK-A1D30D/2020\|EPI_ISL_595048\|20 | ···················· | ···················· | ····················· | ···················· | ···················· | ···················· |
| M | 9394 hCoV-19/England/MILK-A1D31C/2020\|EPI_ISL_595049\|20 | ···················· | ···················· | ····················· | ···················· | ···················· | ···················· |
| M | 9395 hCoV-19/England/MILK-A1D33A/2020\|EPI_ISL_595051\|20 | ···················· | ···················· | ····················· | ···················· | ···················· | ···················· |
| M | 9396 hCoV-19/England/MILK-A1D358/2020\|EPI_ISL_595053\|20 | ···················· | ···················· | ····················· | ···················· | ···················· | ···················· |
| M | 9397 hCoV-19/England/MILK-A1D367/2020\|EPI_ISL_595054\|20 | ···················· | ···················· | ····················· | ···················· | ···················· | ···················· |
| M | 9398 hCoV-19/England/MILK-A1D3C1/2020\|EPI_ISL_595055\|20 | ···················· | ···················· | ····················· | ···················· | ···················· | ···················· |
| M | 9399 hCoV-19/England/MILK-A1D3FE/2020\|EPI_ISL_595056\|20 | ···················· | ·A·················· | ····················· | ···················· | ···················· | ···················· |
| M | 9400 hCoV-19/England/MILK-A1D455/2020\|EPI_ISL_595058\|20 | ···················· | ···················· | ····················· | ···················· | ···················· | ···················· |
| M | 9401 hCoV-19/England/MILK-A1D473/2020\|EPI_ISL_595059\|20 | ···················· | ···················· | ····················· | ···················· | ···················· | ···················· |
| M | 9402 hCoV-19/England/MILK-A1D482/2020\|EPI_ISL_595060\|20 | ···················· | ···················· | ····················· | ···················· | ···················· | ···················· |
| M | 9403 hCoV-19/England/MILK-A1D4A0/2020\|EPI_ISL_595061\|20 | ···················· | ···················· | ····················· | ···················· | ···················· | ···················· |
| M | 9404 hCoV-19/England/MILK-A1D4BF/2020\|EPI_ISL_595062\|20 | ···················· | ···················· | ····················· | ···················· | ···················· | ···················· |
| M | 9405 hCoV-19/England/MILK-A1D4CE/2020\|EPI_ISL_595063\|20 | ···················· | ···················· | ····················· | ···················· | ···················· | ···················· |
| M | 9406 hCoV-19/England/MILK-A1D4DD/2020\|EPI_ISL_595064\|20 | ···················· | ···················· | ····················· | ···················· | ···················· | ···················· |
| M | 9407 hCoV-19/Wales/MILK-A1D4FB/2020\|EPI_ISL_595065\|2020 | ···················· | ···················· | ····················· | ···················· | ···················· | ···················· |
| M | 9408 hCoV-19/England/MILK-A1D507/2020\|EPI_ISL_595066\|20 | ···················· | ···················· | ····················· | ···················· | ···················· | ···················· |
| M | 9409 hCoV-19/England/MILK-A1D552/2020\|EPI_ISL_595068\|20 | ···················· | ···················· | ····················· | ···················· | ···················· | ···················· |
| M | 9410 hCoV-19/England/MILK-A1D67D/2020\|EPI_ISL_595070\|20 | ···················· | ···················· | ····················· | ···················· | ······T············· | ···················· |
| M | 9411 hCoV-19/England/MILK-A1D68C/2020\|EPI_ISL_595071\|20 | ···················· | ···················· | ····················· | ···················· | ···················· | ···················· |
| M | 9412 hCoV-19/England/MILK-A1D6B9/2020\|EPI_ISL_595074\|20 | ···················· | ···················· | ····················· | ···················· | ···················· | ···················· |
| M | 9413 hCoV-19/England/MILK-A1D6D7/2020\|EPI_ISL_595076\|20 | ···················· | ···················· | ····················· | ···················· | ···················· | ···················· |
| M | 9414 hCoV-19/England/MILK-A1D6E6/2020\|EPI_ISL_595077\|20 | ···················· | ···················· | ····················· | ···················· | ···················· | ···················· |
| M | 9415 hCoV-19/England/MILK-A1D701/2020\|EPI_ISL_595078\|20 | ···················· | ···················· | ····················· | ···················· | ···················· | ···················· |
| M | 9416 hCoV-19/England/MILK-A1D710/2020\|EPI_ISL_595079\|20 | ···················· | ···················· | ····················· | ···················· | ···················· | ···················· |
| M | 9417 hCoV-19/Wales/MILK-A1D73E/2020\|EPI_ISL_595080\|2020 | ···················· | ···················· | ····················· | ···················· | ···················· | ···················· |
| M | 9418 hCoV-19/England/MILK-A1D74D/2020\|EPI_ISL_595081\|20 | ···················· | ···················· | ····················· | ···················· | ···················· | ···················· |
| M | 9419 hCoV-19/England/MILK-A1D77A/2020\|EPI_ISL_595082\|20 | ···················· | ···················· | ····················· | ···················· | ······T············· | ···················· |
| M | 9420 hCoV-19/England/MILK-A1D789/2020\|EPI_ISL_595083\|20 | ···················· | ···················· | ····················· | ···················· | ······T············· | ···················· |
| M | 9421 hCoV-19/England/MILK-A1D798/2020\|EPI_ISL_595084\|20 | ···················· | ···················· | ····················· | ···················· | ···················· | ···················· |
| M | 9422 hCoV-19/England/MILK-A1D7B6/2020\|EPI_ISL_595085\|20 | ···················· | ···················· | ····················· | ···················· | ···················· | ···················· |
| M | 9423 hCoV-19/England/MILK-A1D7D4/2020\|EPI_ISL_595087\|20 | ···················· | ···················· | ····················· | ···················· | ···················· | ···················· |
| M | 9424 hCoV-19/England/MILK-A1D7E3/2020\|EPI_ISL_595088\|20 | ···················· | ···················· | ····················· | ···················· | ···················· | ···················· |
| M | 9425 hCoV-19/England/MILK-A1D82C/2020\|EPI_ISL_595091\|20 | ···················· | ···················· | ····················· | ···················· | ···················· | ···················· |
| M | 9426 hCoV-19/England/MILK-A1D886/2020\|EPI_ISL_595093\|20 | ···················· | ···················· | ····················· | ···················· | ···················· | ···················· |
| M | 9427 hCoV-19/England/MILK-A1D895/2020\|EPI_ISL_595094\|20 | ···················· | ···················· | ····················· | ···················· | ···················· | ···················· |
| M | 9428 hCoV-19/England/MILK-A1D8FF/2020\|EPI_ISL_595096\|20 | ···················· | ···················· | ····················· | ···················· | ···················· | ···················· |
| M | 9429 hCoV-19/England/MILK-A1D956/2020\|EPI_ISL_595098\|20 | ···················· | ···················· | ····················· | ···················· | ···················· | ···················· |
| M | 9430 hCoV-19/Russia/SVE-RII-MH1627S/2020\|EPI_ISL_596300 | ···················· | ···················· | ····················· | ···················· | ···················· | ···················· |

| | | | | | | | | |
|---|---|---|---|---|---|---|---|---|
| M | 9431 | hCoV-19/Russia/MOW-RII-MH1980S/2020\|EPI_ISL_596326 | ···················· | ···················· | ···················· | ···················· | ···················· | ···················· |
| M | 9432 | hCoV-19/Russia/MOW-RII-MH1982S/2020\|EPI_ISL_596327 | ···················· | ···················· | ···················· | ···················· | ···················· | ···················· |
| M | 9433 | hCoV-19/Russia/MOW-RII-MH1983S/2020\|EPI_ISL_596328 | ···················· | ···················· | ···················· | ···················· | ···················· | ···················· |
| M | 9434 | hCoV-19/Russia/MOW-RII-MH1984S/2020\|EPI_ISL_596329 | ···················· | ······A············· | ···················· | ···················· | ···················· | ···················· |
| M | 9435 | hCoV-19/Russia/MOW-RII-MH1985S/2020\|EPI_ISL_596330 | ···················· | ······A············· | ···················· | ···················· | ···················· | ···················· |
| M | 9436 | hCoV-19/Russia/MOW-RII-MH1986S/2020\|EPI_ISL_596331 | ···················· | ···················· | ···················· | ···················· | ···················· | ···················· |
| M | 9437 | hCoV-19/Russia/MOW-RII-MH1988S/2020\|EPI_ISL_596332 | ···················· | ···················· | ···················· | ···················· | ···················· | ···················· |
| M | 9438 | hCoV-19/Russia/MOW-RII-MH1989S/2020\|EPI_ISL_596333 | ···················· | ···················· | ···················· | ···················· | ···················· | ···················· |
| M | 9439 | hCoV-19/Russia/MOW-RII-MH2003S/2020\|EPI_ISL_596345 | ···················· | ···················· | ···················· | ···················· | ···················· | ···················· |
| M | 9440 | hCoV-19/Russia/MOW-RII-MH2004S/2020\|EPI_ISL_596346 | ···················· | ···················· | ···················· | ···················· | ···················· | ···················· |
| M | 9441 | hCoV-19/Russia/MOW-RII-MH2005S/2020\|EPI_ISL_596347 | ···················· | ···················· | ···················· | ···················· | ···················· | ···················· |
| M | 9442 | hCoV-19/Russia/MOW-RII-MH2006S/2020\|EPI_ISL_596348 | ···················· | ···················· | ···················· | ···················· | ···················· | ···················· |
| M | 9443 | hCoV-19/Russia/MOW-RII-MH2007S/2020\|EPI_ISL_596349 | ···················· | ···················· | ···················· | ···················· | ···················· | ···················· |
| M | 9444 | hCoV-19/Russia/MOW-RII-MH2008S/2020\|EPI_ISL_596350 | ···················· | ···················· | ···················· | ···················· | ···················· | ···················· |
| M | 9445 | hCoV-19/Russia/MOW-RII-MH2010S/2020\|EPI_ISL_596351 | ···················· | ···················· | ···················· | ···················· | ···················· | ···················· |
| M | 9446 | hCoV-19/Russia/MOW-RII-MH2014S/2020\|EPI_ISL_596352 | ···················· | ···················· | ···················· | ···················· | ···················· | ···················· |
| M | 9447 | hCoV-19/Russia/PRI-RII-MH733S/2020\|EPI_ISL_596353\| | ···················· | ···················· | ···················· | ···················· | ···················· | ···················· |
| M | 9448 | hCoV-19/Russia/PRI-RII-MH744S/2020\|EPI_ISL_596354\| | ···················· | ···················· | ···················· | ···················· | ···················· | ···················· |
| M | 9449 | hCoV-19/Russia/PRI-RII-MH747S/2020\|EPI_ISL_596355\| | ···················· | ···················· | ···················· | ···················· | ···················· | ···················· |
| M | 9450 | hCoV-19/Russia/PRI-RII-MH748S/2020\|EPI_ISL_596356\| | ···················· | ···················· | ···················· | ···················· | ···················· | ···················· |
| M | 9451 | hCoV-19/Russia/SPE-RII-MH939S/2020\|EPI_ISL_596357\| | ···················· | ···················· | ···················· | ···················· | ···················· | ···················· |
| M | 9452 | hCoV-19/Russia/SPE-RII-MH943S/2020\|EPI_ISL_596358\| | ···················· | ···················· | ···················· | ···················· | ···················· | ···················· |
| M | 9453 | hCoV-19/Russia/SPE-RII-MH944S/2020\|EPI_ISL_596359\| | ···················· | ···················· | ···················· | ···················· | ···················· | ···················· |
| M | 9454 | hCoV-19/Russia/SPE-RII-MH945S/2020\|EPI_ISL_596360\| | ···················· | ···················· | ···················· | ···················· | ···················· | ···················· |
| M | 9455 | hCoV-19/Russia/SPE-RII-MH949S/2020\|EPI_ISL_596361\| | ···················· | ···················· | ···················· | ···················· | ···················· | ···················· |
| M | 9456 | hCoV-19/Russia/SPE-RII-MH952S/2020\|EPI_ISL_596362\| | ···················· | ···················· | ···················· | ···················· | ···················· | ···················· |
| M | 9457 | hCoV-19/Russia/SPE-RII-MH953S/2020\|EPI_ISL_596363\| | ···················· | ···················· | ···················· | ···················· | ···················· | ···················· |
| M | 9458 | hCoV-19/Russia/SPE-RII-MH956S/2020\|EPI_ISL_596364\| | ···················· | ···················· | ···················· | ···················· | ···················· | ···················· |
| M | 9459 | hCoV-19/Russia/SPE-RII-MH958S/2020\|EPI_ISL_596365\| | ···················· | ···················· | ···················· | ···················· | ···················· | ···················· |
| M | 9460 | hCoV-19/Russia/SPE-RII-MH960S/2020\|EPI_ISL_596366\| | ···················· | ···················· | ···················· | ···················· | ···················· | ···················· |
| M | 9461 | hCoV-19/Russia/SPE-RII-MH968S/2020\|EPI_ISL_596367\| | ···················· | ···················· | ···················· | ···················· | ···················· | ···················· |
| M | 9462 | hCoV-19/Russia/SPE-RII-MH976S/2020\|EPI_ISL_596368\| | ···················· | ···················· | ···················· | ···················· | ···················· | ···················· |
| M | 9463 | hCoV-19/Russia/SPE-RII-MH977S/2020\|EPI_ISL_596369\| | ···················· | ···················· | ···················· | ···················· | ···················· | ···················· |
| M | 9464 | hCoV-19/Russia/SPE-RII-MH981S/2020\|EPI_ISL_596370\| | ···················· | ···················· | ···················· | ···················· | ···················· | ···················· |
| M | 9465 | hCoV-19/Russia/SPE-RII-MH982S/2020\|EPI_ISL_596371\| | ···················· | ···················· | ···················· | ···················· | ···················· | ···················· |
| M | 9466 | hCoV-19/Russia/SPE-RII-MH983S/2020\|EPI_ISL_596372\| | ···················· | ···················· | ···················· | ···················· | ···················· | ···················· |
| M | 9467 | hCoV-19/Russia/SPE-RII-MH984S/2020\|EPI_ISL_596373\| | ···················· | ···················· | ···················· | ···················· | ···················· | ···················· |
| M | 9468 | hCoV-19/Russia/SPE-RII-MH989S/2020\|EPI_ISL_596374\| | ···················· | ···················· | ···················· | ···················· | ···················· | ···················· |
| M | 9469 | hCoV-19/Russia/SPE-RII-MH995S/2020\|EPI_ISL_596375\| | ···················· | ···················· | ···················· | ···················· | ···················· | ···················· |
| M | 9470 | hCoV-19/Russia/SPE-RII-MH996S/2020\|EPI_ISL_596376\| | ···················· | ···················· | ···················· | ···················· | ···················· | ···················· |
| M | 9471 | hCoV-19/Russia/SPE-RII-MH998S/2020\|EPI_ISL_596377\| | ···················· | ···················· | ···················· | ···················· | ···················· | ···················· |
| M | 9472 | hCoV-19/USA/WA-NIH-WA1/2020\|EPI_ISL_596386\|2020-01 | ···················· | ···················· | ···················· | ···················· | ···················· | ···················· |
| M | 9473 | hCoV-19/England/CAMC-9D6D5E/2020\|EPI_ISL_584324\|20 | ···················· | ·A·················· | ···················· | ···················· | ···················· | ···A················ |
| M | 9474 | hCoV-19/England/CAMC-9D6E2E/2020\|EPI_ISL_584325\|20 | ···················· | ···················· | ···················· | ···················· | ···················· | ···················· |
| M | 9475 | hCoV-19/England/CAMC-9D6E3D/2020\|EPI_ISL_584326\|20 | ···················· | ···················· | ···················· | ···················· | ···················· | ···················· |
| M | 9476 | hCoV-19/England/CAMC-9D6E4C/2020\|EPI_ISL_584327\|20 | ···················· | ···················· | ···················· | ···················· | ···················· | ···················· |
| M | 9477 | hCoV-19/England/CAMC-9D6E5B/2020\|EPI_ISL_584328\|20 | ···················· | ···················· | ···················· | ···················· | ···················· | ···················· |
| M | 9478 | hCoV-19/England/CAMC-9D6E88/2020\|EPI_ISL_584330\|20 | ···················· | ·A·················· | ···················· | ···················· | ···················· | ···A················ |
| M | 9479 | hCoV-19/England/CAMC-9D6E97/2020\|EPI_ISL_584331\|20 | ···················· | ·A·················· | ···················· | ···················· | ···················· | ···A················ |
| M | 9480 | hCoV-19/England/CAMC-9D6EA6/2020\|EPI_ISL_584332\|20 | ···················· | ···················· | ···················· | ···················· | ···················· | ···················· |
| M | 9481 | hCoV-19/England/CAMC-9D6EB5/2020\|EPI_ISL_584333\|20 | ···················· | ···················· | ···················· | ···················· | ···················· | ···················· |
| M | 9482 | hCoV-19/England/CAMC-9D6EE2/2020\|EPI_ISL_584335\|20 | ···················· | ···················· | ···················· | ···················· | ···················· | ···················· |
| M | 9483 | hCoV-19/England/CAMC-9D6F0D/2020\|EPI_ISL_584336\|20 | ···················· | ···················· | ···················· | ···················· | ···················· | ···················· |
| M | 9484 | hCoV-19/England/CAMC-9D6F49/2020\|EPI_ISL_584337\|20 | ···················· | ·A·················· | ···················· | ···················· | ···················· | ···················· |
| M | 9485 | hCoV-19/England/CAMC-9D6F67/2020\|EPI_ISL_584338\|20 | ···················· | ···················· | ···················· | ···················· | ···················· | ···················· |
| M | 9486 | hCoV-19/England/CAMC-9D6FB2/2020\|EPI_ISL_584339\|20 | ···················· | ···················· | ···················· | ···················· | ···················· | ···················· |
| M | 9487 | hCoV-19/England/CAMC-9D7036/2020\|EPI_ISL_584340\|20 | ···················· | ···················· | ···················· | ···················· | ···················· | ···················· |
| M | 9488 | hCoV-19/England/CAMC-9D7090/2020\|EPI_ISL_584343\|20 | ···················· | ···················· | ···················· | ···················· | ···················· | ···················· |
| M | 9489 | hCoV-19/England/CAMC-9D71AC/2020\|EPI_ISL_584351\|20 | ···················· | ···················· | ···················· | ···················· | ···················· | ···················· |
| M | 9490 | hCoV-19/England/CAMC-9D7203/2020\|EPI_ISL_584352\|20 | ···················· | ···················· | ···················· | ···················· | ···················· | ···················· |
| M | 9491 | hCoV-19/England/CAMC-9D7212/2020\|EPI_ISL_584353\|20 | ···················· | ···················· | ···················· | ···················· | ···················· | ···················· |
| M | 9492 | hCoV-19/England/CAMC-9D7230/2020\|EPI_ISL_584355\|20 | ···················· | ···················· | ···················· | ···················· | ···················· | ···················· |
| M | 9493 | hCoV-19/England/CAMC-9D726D/2020\|EPI_ISL_584357\|20 | ···················· | ·A·················· | ···················· | ···················· | ···················· | ···················· |
| M | 9494 | hCoV-19/England/CAMC-9D729A/2020\|EPI_ISL_584358\|20 | ···················· | ····A··············· | ···················· | ···················· | ···················· | ···················· |
| M | 9495 | hCoV-19/England/CAMC-9D72F4/2020\|EPI_ISL_584360\|20 | ···················· | ···················· | ···················· | ···················· | ······T············· | ···················· |
| M | 9496 | hCoV-19/England/CAMC-9D731F/2020\|EPI_ISL_584362\|20 | ···················· | ···················· | ···················· | ···················· | ···················· | ···················· |
| M | 9497 | hCoV-19/England/CAMC-9D736A/2020\|EPI_ISL_584363\|20 | ···················· | ·A·················· | ···················· | ···················· | ···················· | ···A················ |
| M | 9498 | hCoV-19/England/CAMC-9D7379/2020\|EPI_ISL_584364\|20 | ···················· | ·A·················· | ···················· | ···················· | ···················· | ···················· |
| M | 9499 | hCoV-19/England/CAMC-9D7397/2020\|EPI_ISL_584365\|20 | ···················· | ···················· | ···················· | ···················· | ···················· | ···················· |
| M | 9500 | hCoV-19/Wales/PHWC-16DFF4/2020\|EPI_ISL_585653\|2020 | ···················· | ···················· | ···················· | ···················· | ···················· | ···················· |
| M | 9501 | hCoV-19/Wales/PHWC-16E00F/2020\|EPI_ISL_585654\|2020 | ···················· | ···················· | ···················· | ···················· | ···················· | ···················· |
| M | 9503 | hCoV-19/Wales/PHWC-16E02D/2020\|EPI_ISL_585656\|2020 | ···················· | ···················· | ···················· | ···················· | ···················· | ···················· |
| M | 9505 | hCoV-19/Wales/PHWC-16E05A/2020\|EPI_ISL_585659\|2020 | ···················· | ···················· | ···················· | ···················· | ···················· | ···················· |
| M | 9508 | hCoV-19/Wales/PHWC-16E087/2020\|EPI_ISL_585662\|2020 | ···················· | ···················· | ···················· | ···················· | ···················· | ···················· |
| M | 9511 | hCoV-19/Wales/PHWC-16E0B4/2020\|EPI_ISL_585665\|2020 | ···················· | ···················· | ···················· | ···················· | ···················· | ···················· |
| M | 9513 | hCoV-19/Wales/PHWC-16E0D2/2020\|EPI_ISL_585667\|2020 | ···················· | ···················· | ···················· | ···················· | ···················· | ···················· |
| M | 9515 | hCoV-19/Wales/PHWC-16E0F0/2020\|EPI_ISL_585669\|2020 | ···················· | ···················· | ···················· | ···················· | ···················· | ···················· |
| M | 9516 | hCoV-19/Wales/PHWC-16E10C/2020\|EPI_ISL_585670\|2020 | ···················· | ···················· | ···················· | ···················· | ···················· | ···················· |
| M | 9518 | hCoV-19/Wales/PHWC-16E139/2020\|EPI_ISL_585673\|2020 | ···················· | ···················· | ···················· | ···················· | ···················· | ···················· |
| M | 9519 | hCoV-19/Wales/PHWC-16E157/2020\|EPI_ISL_585674\|2020 | ···················· | ···················· | ···················· | ···················· | ···················· | ···················· |

| | | | | | | | | |
|---|---|---|---|---|---|---|---|---|
| M | 9520 | hCoV-19/Wales/PHWC-16E1FD/2020\|EPI_ISL_585683\|2020 | ···················· | ···················· | ···················· | ···················· | ···················· | ···················· |
| M | 9521 | hCoV-19/Wales/PHWC-16E218/2020\|EPI_ISL_585685\|2020 | ···················· | ···················· | ···················· | ···················· | ···················· | ···················· |
| M | 9523 | hCoV-19/Wales/PHWC-16E245/2020\|EPI_ISL_585688\|2020 | ···················· | ···················· | ···················· | ···················· | ···················· | ···················· |
| M | 9524 | hCoV-19/Wales/PHWC-16E290/2020\|EPI_ISL_585693\|2020 | ···················· | ···················· | ···················· | ···················· | ···················· | ···················· |
| M | 9525 | hCoV-19/Wales/PHWC-16E2BE/2020\|EPI_ISL_585695\|2020 | ···················· | ···················· | ···················· | ···················· | ···················· | ···················· |
| M | 9526 | hCoV-19/England/ALDP-A2EF9C/2020\|EPI_ISL_586984\|20 | ···················· | ···················· | ···················· | ···················· | ···················· | ···················· |
| M | 9527 | hCoV-19/England/ALDP-A2E855/2020\|EPI_ISL_586985\|20 | ···················· | ···················· | ···················· | ···················· | ···················· | ···················· |
| M | 9528 | hCoV-19/England/ALDP-A2F1B3/2020\|EPI_ISL_586986\|20 | ···················· | ···················· | ···················· | ···················· | ···················· | ···················· |
| M | 9529 | hCoV-19/England/ALDP-A2F809/2020\|EPI_ISL_586987\|20 | ···················· | ···················· | ···················· | ···················· | ···················· | ···················· |
| M | 9530 | hCoV-19/England/ALDP-A2F362/2020\|EPI_ISL_586988\|20 | ···················· | ···················· | ···················· | ···················· | ···················· | ···················· |
| M | 9531 | hCoV-19/England/ALDP-A2F99D/2020\|EPI_ISL_586989\|20 | ···················· | ···················· | ···················· | ···················· | ···················· | ···················· |
| M | 9532 | hCoV-19/England/ALDP-A2EBC5/2020\|EPI_ISL_586990\|20 | ···················· | ···················· | ···················· | ···················· | ···················· | ···················· |
| M | 9533 | hCoV-19/England/ALDP-A2FC0D/2020\|EPI_ISL_586991\|20 | ···················· | ···················· | ···················· | ···················· | ···················· | ···················· |
| M | 9534 | hCoV-19/England/ALDP-A2FE25/2020\|EPI_ISL_586993\|20 | ···················· | ···················· | ···················· | ···················· | ···················· | ···················· |
| M | 9535 | hCoV-19/England/ALDP-A2E6D3/2020\|EPI_ISL_586994\|20 | ···················· | ···················· | ···················· | ···················· | ···················· | ···················· |
| M | 9536 | hCoV-19/England/ALDP-A2F3DB/2020\|EPI_ISL_586995\|20 | ···················· | ···················· | ···················· | ···················· | ···················· | ···················· |
| M | 9537 | hCoV-19/England/ALDP-A2EE26/2020\|EPI_ISL_586996\|20 | ···················· | ·A·················· | ···················· | ···················· | ···················· | ···A················ |
| M | 9538 | hCoV-19/England/ALDP-A2EAD7/2020\|EPI_ISL_586997\|20 | ···················· | ···················· | ···················· | ···················· | ···················· | ···················· |
| M | 9539 | hCoV-19/England/ALDP-A2FD37/2020\|EPI_ISL_586998\|20 | ···················· | ···················· | ···················· | ···················· | ···················· | ···················· |
| M | 9540 | hCoV-19/England/ALDP-A2F881/2020\|EPI_ISL_586999\|20 | ···················· | ·A·················· | ···················· | ···················· | ···················· | ···A················ |
| M | 9541 | hCoV-19/England/MILK-A17F87/2020\|EPI_ISL_595003\|20 | ···················· | ···················· | ···················· | ···················· | ···················· | ···················· |
| M | 9542 | hCoV-19/England/MILK-A17FA5/2020\|EPI_ISL_595005\|20 | ···················· | ···················· | ···················· | ···················· | ···················· | ···················· |
| M | 9543 | hCoV-19/England/MILK-A17FB4/2020\|EPI_ISL_595006\|20 | ···················· | ···················· | ···················· | ···················· | ···················· | ···················· |
| M | 9544 | hCoV-19/England/MILK-A17FE1/2020\|EPI_ISL_595009\|20 | ···················· | ···················· | ···················· | ···················· | ···················· | ···················· |
| M | 9545 | hCoV-19/England/MILK-A17FF0/2020\|EPI_ISL_595010\|20 | ···················· | ···················· | ···················· | ···················· | ······T············· | ···················· |
| M | 9546 | hCoV-19/Wales/MILK-A1801A/2020\|EPI_ISL_595011\|2020 | ···················· | ···················· | ···················· | ···················· | ···················· | ···················· |
| M | 9547 | hCoV-19/Wales/MILK-A18038/2020\|EPI_ISL_595012\|2020 | ···················· | ···················· | ···················· | ···················· | ···················· | ···················· |
| M | 9548 | hCoV-19/Russia/SPE-RII-MH1628S/2020\|EPI_ISL_596301 | ···················· | ···················· | ···················· | ···················· | ···················· | ···················· |
| M | 9549 | hCoV-19/Russia/SVE-RII-MH1632S/2020\|EPI_ISL_596302 | ···················· | ···················· | ···················· | ···················· | ···················· | ···················· |
| M | 9550 | hCoV-19/Russia/SVE-RII-MH1633S/2020\|EPI_ISL_596303 | ···················· | ···················· | ···················· | ···················· | ···················· | ···················· |
| M | 9551 | hCoV-19/Russia/SVE-RII-MH1634S/2020\|EPI_ISL_596304 | ···················· | ···················· | ···················· | ···················· | ···················· | ···················· |
| M | 9552 | hCoV-19/Russia/SVE-RII-MH1636S/2020\|EPI_ISL_596305 | ···················· | ···················· | ···················· | ···················· | ···················· | ···················· |
| M | 9553 | hCoV-19/Russia/SVE-RII-MH1637S/2020\|EPI_ISL_596306 | ···················· | ···················· | ···················· | ···················· | ···················· | ···················· |
| M | 9554 | hCoV-19/Russia/SVE-RII-MH1638S/2020\|EPI_ISL_596307 | ···················· | ···················· | ···················· | ···················· | ···················· | ···················· |
| M | 9555 | hCoV-19/Russia/SVE-RII-MH1640S/2020\|EPI_ISL_596308 | ···················· | ···················· | ···················· | ···················· | ···················· | ···················· |
| M | 9556 | hCoV-19/Russia/SVE-RII-MH1643S/2020\|EPI_ISL_596309 | ···················· | ···················· | ···················· | ···················· | ···················· | ···················· |
| M | 9557 | hCoV-19/Russia/PER-RII-MH1646S/2020\|EPI_ISL_596310 | ···················· | ···················· | ···················· | ···················· | ···················· | ···················· |
| M | 9558 | hCoV-19/Russia/KK-RII-MH1680S/2020\|EPI_ISL_596311\| | ···················· | ···················· | ···················· | ···················· | ···················· | ···················· |
| M | 9559 | hCoV-19/Russia/KK-RII-MH1681S/2020\|EPI_ISL_596312\| | ···················· | ···················· | ···················· | ···················· | ···················· | ···················· |
| M | 9560 | hCoV-19/Russia/NIZ-RII-MH1682S/2020\|EPI_ISL_596313 | ···················· | ···················· | ···················· | ···················· | ···················· | ···················· |
| M | 9561 | hCoV-19/Russia/NIZ-RII-MH1683S/2020\|EPI_ISL_596314 | ···················· | ···················· | ···················· | ···················· | ···················· | ···················· |
| M | 9562 | hCoV-19/Russia/AD-RII-MH1684S/2020\|EPI_ISL_596315\| | ···················· | ····A··············· | ···················· | ···················· | ···················· | ···················· |
| M | 9563 | hCoV-19/Russia/KDA-RII-MH1685S/2020\|EPI_ISL_596316 | ···················· | ···················· | ···················· | ···················· | ···················· | ···················· |
| M | 9564 | hCoV-19/Russia/KL-RII-MH1686S/2020\|EPI_ISL_596317\| | ···················· | ···················· | ···················· | ···················· | ···················· | ···················· |
| M | 9565 | hCoV-19/Russia/MOW-RII-MH1970S/2020\|EPI_ISL_596318 | ···················· | ···················· | ···················· | ···················· | ···················· | ···················· |
| M | 9566 | hCoV-19/Russia/MOW-RII-MH1971S/2020\|EPI_ISL_596319 | ···················· | ···················· | ···················· | ···················· | ···················· | ···················· |
| M | 9567 | hCoV-19/Russia/MOW-RII-MH1972S/2020\|EPI_ISL_596320 | ···················· | ···················· | ···················· | ···················· | ···················· | ···················· |
| M | 9568 | hCoV-19/Russia/MOW-RII-MH1973S/2020\|EPI_ISL_596321 | ···················· | ···················· | ···················· | ···················· | ···················· | ···················· |
| M | 9569 | hCoV-19/Russia/MOW-RII-MH1974S/2020\|EPI_ISL_596322 | ···················· | ···················· | ···················· | ···················· | ···················· | ···················· |
| M | 9570 | hCoV-19/Russia/MOW-RII-MH1975S/2020\|EPI_ISL_596323 | ···················· | ···················· | ···················· | ···················· | ···················· | ···················· |
| M | 9571 | hCoV-19/Russia/MOW-RII-MH1976S/2020\|EPI_ISL_596324 | ···················· | ···················· | ···················· | ···················· | ···················· | ···················· |
| M | 9572 | hCoV-19/Russia/MOW-RII-MH1979S/2020\|EPI_ISL_596325 | ···················· | ···················· | ···················· | ···················· | ···················· | ···················· |
| M | 9573 | hCoV-19/Russia/MOW-RII-MH1990S/2020\|EPI_ISL_596334 | ···················· | ···················· | ···················· | ···················· | ···················· | ···················· |
| M | 9574 | hCoV-19/Russia/MOW-RII-MH1991S/2020\|EPI_ISL_596335 | ···················· | ···················· | ···················· | ···················· | ···················· | ···················· |
| M | 9575 | hCoV-19/Russia/MOW-RII-MH1992S/2020\|EPI_ISL_596336 | ···················· | ···················· | ···················· | ···················· | ···················· | ···················· |
| M | 9576 | hCoV-19/Russia/MOW-RII-MH1993S/2020\|EPI_ISL_596337 | ···················· | ···················· | ···················· | ···················· | ···················· | ···················· |
| M | 9577 | hCoV-19/Russia/MOW-RII-MH1994S/2020\|EPI_ISL_596338 | ···················· | ···················· | ···················· | ···················· | ···················· | ···················· |
| M | 9578 | hCoV-19/Russia/MOW-RII-MH1995S/2020\|EPI_ISL_596339 | ···················· | ···················· | ···················· | ···················· | ···················· | ···················· |
| M | 9579 | hCoV-19/Russia/MOW-RII-MH1996S/2020\|EPI_ISL_596340 | ···················· | ···················· | ···················· | ···················· | ···················· | ···················· |
| M | 9580 | hCoV-19/Russia/MOW-RII-MH1997S/2020\|EPI_ISL_596341 | ···················· | ···················· | ···················· | ···················· | ···················· | ···················· |
| M | 9581 | hCoV-19/Russia/MOW-RII-MH1998S/2020\|EPI_ISL_596342 | ···················· | ···················· | ···················· | ···················· | ···················· | ···················· |
| M | 9582 | hCoV-19/Russia/MOW-RII-MH1999S/2020\|EPI_ISL_596343 | ···················· | ···················· | ···················· | ···················· | ···················· | ···················· |
| M | 9583 | hCoV-19/Russia/MOW-RII-MH2001S/2020\|EPI_ISL_596344 | ···················· | ···················· | ···················· | ···················· | ···················· | ···················· |
| N | 1 | hCoV-19/Scotland/QEUH-A0B575/2020\|EPI_ISL_599216\|2 | ···················· | ···················· | ···················· | ···················· | ···················· | ···················· |
| N | 2 | hCoV-19/England/QEUH-A0B44B/2020\|EPI_ISL_599217\|20 | ···················· | ···················· | ···················· | ···················· | ···················· | ···················· |
| N | 3 | hCoV-19/England/QEUH-A0C073/2020\|EPI_ISL_599218\|20 | ···················· | ···················· | ···················· | ···················· | ···················· | ···················· |
| N | 4 | hCoV-19/England/QEUH-A0C8E4/2020\|EPI_ISL_599219\|20 | ···················· | ···················· | ···················· | ···················· | ···················· | ···················· |
| N | 5 | hCoV-19/Scotland/QEUH-A0BD6E/2020\|EPI_ISL_599220\|2 | ···················· | ···················· | ···················· | ···················· | ···················· | ···················· |
| N | 6 | hCoV-19/England/QEUH-A0BFB3/2020\|EPI_ISL_599222\|20 | ···················· | ···················· | ···················· | ···················· | ···················· | ···················· |
| N | 7 | hCoV-19/Scotland/QEUH-A0B4D2/2020\|EPI_ISL_599223\|2 | ···················· | ···················· | ···················· | ···················· | ···················· | ···················· |
| N | 8 | hCoV-19/England/QEUH-A0B3D5/2020\|EPI_ISL_599224\|20 | ···················· | ···················· | ···················· | ···················· | ···················· | ···················· |
| N | 9 | hCoV-19/Scotland/QEUH-A0ADF6/2020\|EPI_ISL_599225\|2 | ···················· | ···················· | ···················· | ···················· | ···················· | ···················· |
| N | 10 | hCoV-19/England/QEUH-A0B1DB/2020\|EPI_ISL_599226\|20 | ···················· | ···················· | ···················· | ···················· | ···················· | ···················· |
| N | 11 | hCoV-19/Scotland/QEUH-A0AC9F/2020\|EPI_ISL_599227\|2 | ···················· | ···················· | ···················· | ···················· | ···················· | ···················· |
| N | 12 | hCoV-19/England/QEUH-A0B618/2020\|EPI_ISL_599229\|20 | ···················· | ···················· | ···················· | ···················· | ···················· | ···················· |
| N | 13 | hCoV-19/England/QEUH-A0B311/2020\|EPI_ISL_599230\|20 | ···················· | ···················· | ···················· | ···················· | ···················· | ···················· |
| N | 14 | hCoV-19/Scotland/QEUH-A0BF1D/2020\|EPI_ISL_599231\|2 | ···················· | ···················· | ···················· | ···················· | ···················· | ···················· |
| N | 15 | hCoV-19/England/QEUH-A0B469/2020\|EPI_ISL_599232\|20 | ···················· | ···················· | ···················· | ···················· | ···················· | ···················· |
| N | 16 | hCoV-19/Scotland/QEUH-A0B3E4/2020\|EPI_ISL_599233\|2 | ···················· | ···················· | ···················· | ···················· | ···················· | ···················· |
| N | 17 | hCoV-19/England/QEUH-A0AE99/2020\|EPI_ISL_599234\|20 | ···················· | ···················· | ···················· | ···················· | ···················· | ···················· |

| | | | | | | | |
|---|---|---|---|---|---|---|---|
| N | 18 hCoV-19/Scotland/QEUH-A0B8E5/2020\|EPI_ISL_599235\|2 | ···················· | ···················· | ····················· | ···················· | ···················· | ···················· |
| N | 19 hCoV-19/Scotland/QEUH-A0B4A5/2020\|EPI_ISL_599236\|2 | ···················· | ···················· | ····················· | ···················· | ···················· | ···················· |
| N | 20 hCoV-19/Scotland/QEUH-A0B86D/2020\|EPI_ISL_599237\|2 | ···················· | ···················· | ····················· | ···················· | ···················· | ···················· |
| N | 21 hCoV-19/Scotland/QEUH-A0AD50/2020\|EPI_ISL_599238\|2 | ···················· | ···················· | ····················· | ···················· | ···················· | ···················· |
| N | 22 hCoV-19/Scotland/QEUH-A0B6BE/2020\|EPI_ISL_599239\|2 | ···················· | ···················· | ····················· | ···················· | ···················· | ···················· |
| N | 23 hCoV-19/England/QEUH-A0BDF5/2020\|EPI_ISL_599240\|20 | ···················· | ···················· | ····················· | ···················· | ···················· | ···················· |
| N | 24 hCoV-19/Scotland/QEUH-A0B78E/2020\|EPI_ISL_599241\|2 | ···················· | ···················· | ····················· | ···················· | ···················· | ···················· |
| N | 25 hCoV-19/England/QEUH-A0B153/2020\|EPI_ISL_599242\|20 | ···················· | ···················· | ····················· | ···················· | ···················· | ···················· |
| N | 26 hCoV-19/Scotland/QEUH-A0C0BF/2020\|EPI_ISL_599243\|2 | ···················· | ···················· | ····················· | ···················· | ···················· | ···················· |
| N | 27 hCoV-19/England/QEUH-A0BBEC/2020\|EPI_ISL_599244\|20 | ···················· | ···················· | ····················· | ···················· | ···················· | ···················· |
| N | 28 hCoV-19/England/QEUH-A0B663/2020\|EPI_ISL_599245\|20 | ···················· | ···················· | ····················· | ···················· | ···················· | ···················· |
| N | 29 hCoV-19/England/QEUH-A0C83F/2020\|EPI_ISL_599246\|20 | ···················· | ···················· | ····················· | ···················· | ···················· | ···················· |
| N | 30 hCoV-19/Scotland/QEUH-A0B1F9/2020\|EPI_ISL_599247\|2 | ···················· | ···················· | ····················· | ···················· | ···················· | ···················· |
| N | 31 hCoV-19/Scotland/QEUH-A0B566/2020\|EPI_ISL_599248\|2 | ···················· | ···················· | ····················· | ···················· | ···················· | ···················· |
| N | 32 hCoV-19/Scotland/QEUH-A0B8B8/2020\|EPI_ISL_599249\|2 | ···················· | ···················· | ····················· | ···················· | ···················· | ···················· |
| N | 33 hCoV-19/England/QEUH-A0C88A/2020\|EPI_ISL_599250\|20 | ···················· | ···················· | ····················· | ···················· | ···················· | ···················· |
| N | 34 hCoV-19/England/QEUH-A0AF69/2020\|EPI_ISL_599251\|20 | ···················· | ···················· | ····················· | ···················· | ···················· | ···················· |
| N | 35 hCoV-19/England/QEUH-A0BE7A/2020\|EPI_ISL_599252\|20 | ···················· | ···················· | ····················· | ···················· | ···················· | ···················· |
| N | 36 hCoV-19/England/QEUH-A0B645/2020\|EPI_ISL_599254\|20 | ···················· | ···················· | ····················· | ···················· | ···················· | ···················· |
| N | 37 hCoV-19/Scotland/QEUH-A0B77F/2020\|EPI_ISL_599255\|2 | ···················· | ···················· | ····················· | ···················· | ···················· | ···················· |
| N | 38 hCoV-19/Scotland/QEUH-A0ADD8/2020\|EPI_ISL_599256\|2 | ···················· | ···················· | ····················· | ···················· | ···················· | ···················· |
| N | 39 hCoV-19/Scotland/QEUH-A0B302/2020\|EPI_ISL_599257\|2 | ···················· | ···················· | ····················· | ···················· | ···················· | ···················· |
| N | 40 hCoV-19/England/QEUH-A0B01A/2020\|EPI_ISL_599258\|20 | ···················· | ···················· | ····················· | ···················· | ···················· | ···················· |
| N | 41 hCoV-19/England/QEUH-A0B4E1/2020\|EPI_ISL_599259\|20 | ···················· | ···················· | ····················· | ···················· | ···················· | ···················· |
| N | 42 hCoV-19/England/QEUH-A0C87B/2020\|EPI_ISL_599280\|20 | ···················· | ···················· | ····················· | ···················· | ···················· | ···················· |
| N | 43 hCoV-19/England/QEUH-A0B3F3/2020\|EPI_ISL_599282\|20 | ···················· | ·A·················· | ····················· | ···················· | ···················· | ···················· |
| N | 44 hCoV-19/England/QEUH-A0B52A/2020\|EPI_ISL_599283\|20 | ···················· | ···················· | ····················· | ···················· | ···················· | ···················· |
| N | 45 hCoV-19/England/QEUH-A0C3A7/2020\|EPI_ISL_599284\|20 | ···················· | ···················· | ····················· | ···················· | ···················· | ···················· |
| N | 46 hCoV-19/England/QEUH-A0B0CF/2020\|EPI_ISL_599285\|20 | ···················· | ···················· | ····················· | ···················· | ···················· | ···················· |
| N | 47 hCoV-19/Scotland/QEUH-A0BD5F/2020\|EPI_ISL_599286\|2 | ···················· | ···················· | ····················· | ···················· | ···················· | ···················· |
| N | 48 hCoV-19/England/QEUH-A0AC08/2020\|EPI_ISL_599287\|20 | ···················· | ···················· | ····················· | ···················· | ···················· | ···················· |
| N | 49 hCoV-19/England/QEUH-A0BF77/2020\|EPI_ISL_599288\|20 | ···················· | ···················· | ····················· | ···················· | ···················· | ···················· |
| N | 50 hCoV-19/England/QEUH-A0AF96/2020\|EPI_ISL_599289\|20 | ···················· | ···················· | ····················· | ···················· | ···················· | ···················· |
| N | 51 hCoV-19/England/QEUH-A0ADAB/2020\|EPI_ISL_599290\|20 | ···················· | ···················· | ····················· | ···················· | ·····T·············· | ···················· |
| N | 52 hCoV-19/Scotland/QEUH-A0BF0E/2020\|EPI_ISL_599291\|2 | ···················· | ···················· | ····················· | ···················· | ···················· | ···················· |
| N | 53 hCoV-19/England/QEUH-A0AA95/2020\|EPI_ISL_599292\|20 | ···················· | ···················· | ····················· | ···················· | ···················· | ···················· |
| N | 54 hCoV-19/Scotland/QEUH-A0AE5D/2020\|EPI_ISL_599293\|2 | ···················· | ···················· | ····················· | ···················· | ···················· | ···················· |
| N | 55 hCoV-19/Scotland/QEUH-A0B2D8/2020\|EPI_ISL_599294\|2 | ···················· | ···················· | ····················· | ···················· | ···················· | ···················· |
| N | 56 hCoV-19/Scotland/QEUH-A0BEE3/2020\|EPI_ISL_599295\|2 | ···················· | ···················· | ····················· | ···················· | ···················· | ···················· |
| N | 57 hCoV-19/England/QEUH-A0BE98/2020\|EPI_ISL_599296\|20 | ···················· | ···················· | ····················· | ···················· | ···················· | ···················· |
| N | 58 hCoV-19/England/QEUH-A0BE4D/2020\|EPI_ISL_599297\|20 | ···················· | ···················· | ····················· | ···················· | ···················· | ···················· |
| N | 59 hCoV-19/Wales/QEUH-A0B627/2020\|EPI_ISL_599298\|2020 | ···················· | ···················· | ····················· | ···················· | ···················· | ···················· |
| N | 60 hCoV-19/Scotland/QEUH-A0B7BB/2020\|EPI_ISL_599299\|2 | ···················· | ···················· | ····················· | ···················· | ···················· | ···················· |
| N | 61 hCoV-19/USA/MN-MDH-1868/2020\|EPI_ISL_603003\|2020-0 | ···················· | ···················· | ····················· | ···················· | ···················· | ···················· |
| N | 62 hCoV-19/USA/MN-MDH-1869/2020\|EPI_ISL_603004\|2020-0 | ···················· | ···················· | ····················· | ···················· | ···················· | ···················· |
| N | 63 hCoV-19/USA/MN-MDH-1870/2020\|EPI_ISL_603005\|2020-1 | ···················· | ···················· | ····················· | ···················· | ···················· | ···················· |
| N | 64 hCoV-19/USA/MN-MDH-1872/2020\|EPI_ISL_603007\|2020-1 | ···················· | ·A·················· | ····················· | ···················· | ···················· | ···················· |
| N | 65 hCoV-19/USA/MN-MDH-1873/2020\|EPI_ISL_603008\|2020-1 | ···················· | ···················· | ····················· | ···················· | ···················· | ···················· |
| N | 66 hCoV-19/USA/MN-MDH-1874/2020\|EPI_ISL_603009\|2020-1 | ···················· | ···················· | ····················· | ···················· | ···················· | ···················· |
| N | 67 hCoV-19/Brazil/SP-433/2020\|EPI_ISL_603030\|2020-07- | ···················· | ···················· | ····················· | ···················· | ···················· | ···················· |
| N | 68 hCoV-19/Brazil/SP-434/2020\|EPI_ISL_603031\|2020-07- | ···················· | ···················· | ····················· | ···················· | ···················· | ···················· |
| N | 69 hCoV-19/Brazil/SP-435/2020\|EPI_ISL_603032\|2020-07- | ···················· | ···················· | ····················· | ···················· | ···················· | ···················· |
| N | 70 hCoV-19/Brazil/SP-436/2020\|EPI_ISL_603033\|2020-07- | ···················· | ···················· | ····················· | ···················· | ···················· | ···················· |
| N | 71 hCoV-19/Brazil/SP-437/2020\|EPI_ISL_603034\|2020-07- | ···················· | ···················· | ····················· | ···················· | ···················· | ···················· |
| N | 72 hCoV-19/Brazil/SP-438/2020\|EPI_ISL_603035\|2020-07- | ···················· | ···················· | ····················· | ···················· | ···················· | ···················· |
| N | 73 hCoV-19/Brazil/SP-439/2020\|EPI_ISL_603036\|2020-07- | ···················· | ···················· | ····················· | ···················· | ···················· | ···················· |
| N | 74 hCoV-19/Brazil/SP-440/2020\|EPI_ISL_603037\|2020-07- | ···················· | ···················· | ····················· | ···················· | ···········T········ | ···················· |
| N | 75 hCoV-19/Brazil/SP-441/2020\|EPI_ISL_603038\|2020-07- | ···················· | ···················· | ····················· | ···················· | ···················· | ··············A····· |
| N | 76 hCoV-19/Brazil/SP-442/2020\|EPI_ISL_603039\|2020-07- | ···················· | ···················· | ····················· | ···················· | ···················· | ···················· |
| N | 77 hCoV-19/Australia/VIC13521/2020\|EPI_ISL_603040\|202 | ···················· | ···················· | ····················· | ···················· | ···················· | ···················· |
| N | 78 hCoV-19/Australia/VIC15398/2020\|EPI_ISL_603047\|202 | ···················· | ···················· | ····················· | ···················· | ···················· | ···················· |
| N | 80 hCoV-19/USA/UT-UPHL-00022/2020\|EPI_ISL_603050\|2020 | ···················· | ···················· | ····················· | ···················· | ···················· | ···················· |
| N | 81 hCoV-19/USA/UT-UPHL-00023/2020\|EPI_ISL_603051\|2020 | ···················· | ···················· | ····················· | ···················· | ···················· | ···················· |
| N | 82 hCoV-19/USA/UT-UPHL-00025/2020\|EPI_ISL_603052\|2020 | ···················· | ···················· | ····················· | ···················· | ···················· | ···················· |
| N | 83 hCoV-19/USA/CA-QDX-2002/2020\|EPI_ISL_604300\|2020-0 | ···················· | ···················· | ····················· | ···················· | ···················· | ···················· |
| N | 84 hCoV-19/USA/CA-QDX-2003/2020\|EPI_ISL_604301\|2020-0 | ···················· | ···················· | ····················· | ···················· | ···················· | ···················· |
| N | 85 hCoV-19/USA/CA-QDX-2005/2020\|EPI_ISL_604302\|2020-0 | ···················· | ···················· | ····················· | ···················· | ···················· | ···················· |
| N | 86 hCoV-19/USA/CA-QDX-2006/2020\|EPI_ISL_604303\|2020-0 | ···················· | ···················· | ····················· | ···················· | ···················· | ···················· |
| N | 87 hCoV-19/USA/CA-QDX-2007/2020\|EPI_ISL_604304\|2020-0 | ···················· | ···················· | ····················· | ···················· | ···················· | ···················· |
| N | 88 hCoV-19/USA/CA-QDX-2010/2020\|EPI_ISL_604305\|2020-0 | ···················· | ···················· | ····················· | ···················· | ···················· | ···················· |
| N | 89 hCoV-19/USA/CA-QDX-2012/2020\|EPI_ISL_604306\|2020-0 | ···················· | ···················· | ····················· | ···················· | ···················· | ···················· |
| N | 90 hCoV-19/USA/CA-QDX-2013/2020\|EPI_ISL_604307\|2020-0 | ···················· | ···················· | ····················· | ···················· | ···················· | ···················· |
| N | 91 hCoV-19/USA/CA-QDX-2019/2020\|EPI_ISL_604308\|2020-0 | ···················· | ···················· | ····················· | ···················· | ···················· | ···················· |
| N | 92 hCoV-19/USA/CA-QDX-2021/2020\|EPI_ISL_604309\|2020-0 | ···················· | ···················· | ····················· | ···················· | ···················· | ···················· |
| N | 93 hCoV-19/USA/CA-QDX-2022/2020\|EPI_ISL_604310\|2020-0 | ···················· | ···················· | ····················· | ···················· | ···················· | ···················· |
| N | 94 hCoV-19/USA/CA-QDX-2027/2020\|EPI_ISL_604311\|2020-0 | ···················· | ···················· | ····················· | ···················· | ···················· | ···················· |
| N | 95 hCoV-19/USA/CA-QDX-2029/2020\|EPI_ISL_604312\|2020-0 | ···················· | ···················· | ····················· | ···················· | ···················· | ···················· |
| N | 96 hCoV-19/USA/CA-QDX-2031/2020\|EPI_ISL_604313\|2020-0 | ···················· | ···················· | ····················· | ···················· | ···················· | ···················· |
| N | 97 hCoV-19/USA/CA-QDX-2032/2020\|EPI_ISL_604314\|2020-0 | ···················· | ···················· | ····················· | ···················· | ···················· | ···················· |
| N | 98 hCoV-19/USA/CA-QDX-2033/2020\|EPI_ISL_604315\|2020-0 | ···················· | ···················· | ····················· | ···················· | ···················· | ···················· |

N 99 hCoV-19/USA/CA-QDX-2035/2020|EPI_ISL_604316|2020-0 ···················· ···················· ···················· ···················· ···················· ····················
N 100 hCoV-19/USA/CA-QDX-2037/2020|EPI_ISL_604317|2020-0 ···················· ···················· ···················· ···················· ···················· ····················
N 101 hCoV-19/USA/CA-QDX-2038/2020|EPI_ISL_604318|2020-0 ···················· ···················· ···················· ···················· ···················· ····················
N 102 hCoV-19/USA/CA-QDX-2039/2020|EPI_ISL_604319|2020-0 ···················· ···················· ···················· ···················· ···················· ····················
N 103 hCoV-19/USA/CA-QDX-2047/2020|EPI_ISL_604320|2020-0 ···················· ···················· ···················· ···················· ···················· ····················
N 104 hCoV-19/USA/CA-QDX-2052/2020|EPI_ISL_604321|2020-0 ···················· ···················· ···················· ···················· ···················· ····················
N 105 hCoV-19/USA/CA-QDX-2066/2020|EPI_ISL_604322|2020-0 ···················· ···················· ···················· ···················· ···················· ····················
N 106 hCoV-19/USA/CA-QDX-2068/2020|EPI_ISL_604323|2020-0 ···················· ···················· ···················· ···················· ···················· ····················
N 107 hCoV-19/USA/CA-QDX-2071/2020|EPI_ISL_604324|2020-0 ···················· ···················· ···················· ···················· ···················· ····················
N 108 hCoV-19/USA/CA-QDX-2072/2020|EPI_ISL_604325|2020-0 ···················· ···················· ···················· ···················· ···················· ····················
N 109 hCoV-19/USA/FL-QDX-2018/2020|EPI_ISL_604326|2020-0 ···················· ···················· ···················· ···················· ···················· ····················
N 110 hCoV-19/USA/FL-QDX-2057/2020|EPI_ISL_604327|2020-0 ···················· ···················· ···················· ···················· ···················· ····················
N 111 hCoV-19/USA/ID-QDX-2017/2020|EPI_ISL_604328|2020-0 ···················· ···················· ···················· ···················· ···················· ····················
N 112 hCoV-19/USA/ID-QDX-2045/2020|EPI_ISL_604329|2020-0 ···················· ···················· ···················· ···················· ···················· ····················
N 113 hCoV-19/USA/IN-QDX-2069/2020|EPI_ISL_604334|2020-0 ···················· ···················· ···················· ···················· ···················· ····················
N 114 hCoV-19/USA/LA-QDX-1992/2020|EPI_ISL_604335|2020-0 ···················· ···················· ···················· ···················· ···················· ····················
N 115 hCoV-19/USA/LA-QDX-1997/2020|EPI_ISL_604336|2020-0 ···················· ···················· ···················· ···················· ···················· ····················
N 116 hCoV-19/USA/LA-QDX-2000/2020|EPI_ISL_604338|2020-0 ···················· ···················· ···················· ···················· ···················· ····················
N 117 hCoV-19/USA/LA-QDX-2026/2020|EPI_ISL_604339|2020-0 ···················· ···················· ···················· ···················· ···················· ····················
N 118 hCoV-19/USA/NY-QDX-1999/2020|EPI_ISL_604360|2020-0 ···················· ···················· ···················· ···················· ···················· ····················
N 119 hCoV-19/USA/NY-QDX-2008/2020|EPI_ISL_604361|2020-0 ···················· ···················· ···················· ···················· ···················· ····················
N 120 hCoV-19/USA/NY-QDX-2009/2020|EPI_ISL_604362|2020-0 ···················· ···················· ···················· ···················· ···················· ····················
N 121 hCoV-19/USA/NY-QDX-2034/2020|EPI_ISL_604363|2020-0 ···················· ···················· ···················· ···················· ···················· ····················
N 122 hCoV-19/USA/NY-QDX-2043/2020|EPI_ISL_604364|2020-0 ···················· ···················· ···················· ···················· ···················· ····················
N 123 hCoV-19/USA/NY-QDX-2044/2020|EPI_ISL_604365|2020-0 ···················· ···················· ···················· ···················· ···················· ····················
N 124 hCoV-19/USA/NY-QDX-2046/2020|EPI_ISL_604366|2020-0 ···················· ···················· ···················· ···················· ···················· ····················
N 125 hCoV-19/USA/NY-QDX-2051/2020|EPI_ISL_604367|2020-0 ···················· ···················· ···················· ···················· ···················· ····················
N 126 hCoV-19/USA/NY-QDX-2058/2020|EPI_ISL_604368|2020-0 ···················· ···················· ···················· ···················· ···················· ····················
N 127 hCoV-19/USA/NY-QDX-2060/2020|EPI_ISL_604369|2020-0 ···················· ···················· ···················· ···················· ···················· ····················
N 128 hCoV-19/USA/NY-QDX-2063/2020|EPI_ISL_604370|2020-0 ···················· ···················· ···················· ···················· ···················· ····················
N 129 hCoV-19/USA/NY-QDX-2067/2020|EPI_ISL_604371|2020-0 ···················· ···················· ···················· ···················· ···················· ····················
N 130 hCoV-19/USA/OH-QDX-2050/2020|EPI_ISL_604372|2020-0 ···················· ···················· ···················· ···················· ···················· ····················
N 131 hCoV-19/USA/PA-QDX-2001/2020|EPI_ISL_604373|2020-0 ···················· ···················· ···················· ···················· ···················· ····················
N 132 hCoV-19/USA/PA-QDX-2014/2020|EPI_ISL_604374|2020-0 ···················· ···················· ···················· ···················· ···················· ····················
N 133 hCoV-19/USA/PA-QDX-2049/2020|EPI_ISL_604375|2020-0 ···················· ···················· ···················· ···················· ···················· ····················
N 134 hCoV-19/USA/TX-QDX-2025/2020|EPI_ISL_604376|2020-0 ···················· ···················· ···················· ···················· ···················· ····················
N 135 hCoV-19/USA/TX-QDX-2048/2020|EPI_ISL_604377|2020-0 ···················· ···················· ···················· ···················· ···················· ····················
N 136 hCoV-19/USA/UT-QDX-1989/2020|EPI_ISL_604378|2020-0 ···················· ···················· ···················· ···················· ···················· ····················
N 137 hCoV-19/USA/UT-QDX-2004/2020|EPI_ISL_604379|2020-0 ···················· ···················· ···················· ···················· ···················· ····················
N 138 hCoV-19/USA/UT-QDX-2016/2020|EPI_ISL_604380|2020-0 ···················· ···················· ···················· ···················· ···················· ····················
N 139 hCoV-19/USA/UT-QDX-2020/2020|EPI_ISL_604381|2020-0 ···················· ···················· ···················· ···················· ···················· ····················
N 140 hCoV-19/USA/UT-QDX-2024/2020|EPI_ISL_604382|2020-0 ···················· ···················· ···················· ···················· ···················· ····················
N 141 hCoV-19/USA/UT-QDX-2028/2020|EPI_ISL_604383|2020-0 ···················· ···················· ···················· ···················· ···················· ····················
N 142 hCoV-19/USA/UT-QDX-2065/2020|EPI_ISL_604384|2020-0 ···················· ···················· ···················· ···················· ···················· ····················
N 143 hCoV-19/USA/UT-QDX-2070/2020|EPI_ISL_604385|2020-0 ···················· ···················· ···················· ···················· ···················· ····················
N 144 hCoV-19/USA/AR-QDX-2132/2020|EPI_ISL_604386|2020-0 ···················· ···················· ···················· ···················· ···················· ····················
N 145 hCoV-19/USA/AZ-QDX-2118/2020|EPI_ISL_604387|2020-0 ···················· ···················· ···················· ···················· ···················· ····················
N 146 hCoV-19/USA/AZ-QDX-2135/2020|EPI_ISL_604388|2020-0 ···················· ···················· ···················· ···················· ···················· ····················
N 147 hCoV-19/USA/AZ-QDX-2154/2020|EPI_ISL_604389|2020-0 ···················· ···················· ···················· ···················· ···················· ····················
N 148 hCoV-19/USA/AZ-QDX-2156/2020|EPI_ISL_604390|2020-0 ···················· ···················· ···················· ···················· ···················· ····················
N 149 hCoV-19/USA/AZ-QDX-2163/2020|EPI_ISL_604391|2020-0 ···················· ···················· ···················· ···················· ···················· ····················
N 150 hCoV-19/USA/AZ-QDX-2165/2020|EPI_ISL_604392|2020-0 ···················· ···················· ···················· ···················· ···················· ····················
N 151 hCoV-19/USA/CA-QDX-2074/2020|EPI_ISL_604393|2020-0 ···················· ···················· ···················· ···················· ···················· ····················
N 152 hCoV-19/USA/CA-QDX-2075/2020|EPI_ISL_604394|2020-0 ···················· ···················· ···················· ···················· ···················· ····················
N 153 hCoV-19/USA/CA-QDX-2077/2020|EPI_ISL_604395|2020-0 ···················· ···················· ···················· ···················· ···················· ····················
N 154 hCoV-19/USA/CA-QDX-2078/2020|EPI_ISL_604396|2020-0 ···················· ···················· ···················· ···················· ···················· ····················
N 155 hCoV-19/USA/CA-QDX-2080/2020|EPI_ISL_604397|2020-0 ···················· ···················· ···················· ···················· ···················· ····················
N 156 hCoV-19/USA/CA-QDX-2083/2020|EPI_ISL_604398|2020-0 ···················· ···················· ···················· ···················· ···················· ····················
N 157 hCoV-19/USA/CA-QDX-2084/2020|EPI_ISL_604399|2020-0 ···················· ···················· ···················· ···················· ···················· ····················
N 171 hCoV-19/USA/WI-UW-1740/2020|EPI_ISL_605613|2020-09 ···················· ···················· ···················· ···················· ···················· ····················
N 172 hCoV-19/USA/WI-UW-1741/2020|EPI_ISL_605614|2020-09 ···················· ·A·················· ···················· ···················· ···················· ····················
N 173 hCoV-19/USA/WI-UW-1742/2020|EPI_ISL_605615|2020-09 ···················· ·············A······ ···················· ···················· ···················· ····················
N 174 hCoV-19/USA/WI-UW-1743/2020|EPI_ISL_605616|2020-09 ···················· ···················· ···················· ···················· ···················· ····················
N 175 hCoV-19/USA/WI-UW-1744/2020|EPI_ISL_605617|2020-09 ···················· ···················· ···················· ···················· ···················· ····················
N 176 hCoV-19/USA/WI-UW-1745/2020|EPI_ISL_605618|2020-09 ···················· ·············A······ ···················· ···················· ···················· ····················
N 178 hCoV-19/USA/WI-UW-1747/2020|EPI_ISL_605620|2020-09 ···················· ···················· ···················· ···················· ···················· ····················
N 179 hCoV-19/USA/WI-UW-1748/2020|EPI_ISL_605621|2020-09 ···················· ···················· ···················· ···················· ···················· ····················
N 180 hCoV-19/USA/WI-UW-1749/2020|EPI_ISL_605622|2020-09 ···················· ···················· ···················· ···················· ···················· ····················
N 181 hCoV-19/USA/WI-UW-1751/2020|EPI_ISL_605624|2020-09 ···················· ···················· ···················· ···················· ···················· ····················
N 182 hCoV-19/USA/WI-UW-1752/2020|EPI_ISL_605625|2020-09 ···················· ···················· ···················· ···················· ···················· ····················
N 183 hCoV-19/USA/WI-UW-1753/2020|EPI_ISL_605626|2020-09 ···················· ···················· ···················· ···················· ···················· ····················
N 184 hCoV-19/USA/WI-UW-1754/2020|EPI_ISL_605627|2020-09 ···················· ···················· ···················· ···················· ···················· ····················
N 185 hCoV-19/USA/WI-UW-1755/2020|EPI_ISL_605628|2020-09 ···················· ···················· ···················· ···················· ···················· ····················
N 186 hCoV-19/USA/WI-UW-1756/2020|EPI_ISL_605629|2020-09 ···················· ···················· ···················· ···················· ···················· ····················
N 187 hCoV-19/USA/WI-UW-1757/2020|EPI_ISL_605630|2020-09 ···················· ···················· ···················· ···················· ···················· ····················
N 188 hCoV-19/USA/WI-UW-1758/2020|EPI_ISL_605631|2020-09 ···················· ·············A······ ···················· ···················· ···················· ····················
N 189 hCoV-19/USA/WI-UW-1759/2020|EPI_ISL_605632|2020-09 ···················· ···················· ···················· ···················· ···················· ····················
N 190 hCoV-19/USA/WI-UW-1760/2020|EPI_ISL_605633|2020-09 ···················· ···················· ···················· ···················· ···················· ····················
N 191 hCoV-19/USA/WI-UW-1761/2020|EPI_ISL_605634|2020-09 ···················· ···················· ···················· ···················· ···················· ····················
N 192 hCoV-19/USA/WI-UW-1762/2020|EPI_ISL_605635|2020-09 ···················· ···················· ···················· ···················· ···················· ····················

```
N 193 hCoV-19/USA/WI-UW-1763/2020|EPI_ISL_605636|2020-09 ···················· ···················· ····················· ···················· ···················· ····················
N 194 hCoV-19/USA/WI-UW-1764/2020|EPI_ISL_605637|2020-09 ···················· ···················· ····················· ···················· ···················· ····················
N 195 hCoV-19/USA/WI-UW-1765/2020|EPI_ISL_605638|2020-09 ···················· ···················· ····················· ···················· ···················· ····················
N 197 hCoV-19/USA/WI-UW-1769/2020|EPI_ISL_605641|2020-09 ···················· ···················· ····················· ···················· ···················· ····················
N 198 hCoV-19/USA/WI-UW-1770/2020|EPI_ISL_605642|2020-09 ···················· ·············A······ ····················· ···················· ···················· ····················
N 199 hCoV-19/USA/WI-UW-1771/2020|EPI_ISL_605643|2020-09 ···················· ···················· ····················· ···················· ···················· ····················
N 202 hCoV-19/USA/WI-UW-1774/2020|EPI_ISL_605646|2020-09 ···················· ···················· ····················· ···················· ···················· ····················
N 203 hCoV-19/USA/WI-UW-1775/2020|EPI_ISL_605647|2020-09 ···················· ···················· ····················· ···················· ···················· ····················
N 204 hCoV-19/USA/WI-UW-1776/2020|EPI_ISL_605648|2020-09 ···················· ···················· ····················· ···················· ···················· ····················
N 205 hCoV-19/USA/WI-UW-1777/2020|EPI_ISL_605649|2020-09 ···················· ···················· ····················· ···················· ···················· ····················
N 207 hCoV-19/USA/WI-UW-1779/2020|EPI_ISL_605651|2020-09 ···················· ···················· ····················· ···················· ···················· ····················
N 208 hCoV-19/USA/WI-UW-1780/2020|EPI_ISL_605652|2020-09 ···················· ···················· ····················· ···················· ···················· ····················
N 209 hCoV-19/USA/WI-UW-1781/2020|EPI_ISL_605653|2020-09 ···················· ···················· ····················· ···················· ···················· ····················
N 210 hCoV-19/USA/WI-UW-1782/2020|EPI_ISL_605654|2020-09 ···················· ···················· ····················· ···················· ···················· ····················
N 211 hCoV-19/USA/WI-UW-1783/2020|EPI_ISL_605655|2020-09 ···················· ···················· ····················· ···················· ···················· ····················
N 220 hCoV-19/USA/WI-UW-1818/2020|EPI_ISL_605690|2020-09 ···················· ···················· ····················· ···················· ···················· ····················
N 227 hCoV-19/USA/WI-UW-1827/2020|EPI_ISL_605699|2020-09 ···················· ···················· ····················· ···················· ···················· ····················
N 228 hCoV-19/Hong Kong/HKPU-0158/2020|EPI_ISL_610165|20 ···················· ···················· ····················· ···················· ···················· ····················
N 229 hCoV-19/Hong Kong/HKPU-0176/2020|EPI_ISL_610166|20 ···················· ···················· ····················· ···················· ···················· ····················
N 230 hCoV-19/Hong Kong/HKPU-0291/2020|EPI_ISL_610167|20 ···················· ···················· ····················· ···················· ···················· ····················
N 231 hCoV-19/Hong Kong/HKPU-0481/2020|EPI_ISL_610168|20 ···················· ···················· ····················· ···················· ···················· ····················
N 232 hCoV-19/Hong Kong/HKPU-0501/2020|EPI_ISL_610169|20 ···················· ···················· ····················· ···················· ···················· ····················
N 233 hCoV-19/Hong Kong/HKPU-0477/2020|EPI_ISL_610170|20 ···················· ···················· ····················· ···················· ···················· ····················
N 234 hCoV-19/Hong Kong/HKPU-0440/2020|EPI_ISL_610171|20 ···················· ···················· ····················· ···················· ···················· ····················
N 235 hCoV-19/Hong Kong/HKPU-0248/2020|EPI_ISL_610172|20 ···················· ···················· ····················· ···················· ···················· ····················
N 236 hCoV-19/Hong Kong/HKPU-0422/2020|EPI_ISL_610173|20 ···················· ···················· ····················· ···················· ···················· ····················
N 237 hCoV-19/Hong Kong/HKPU-0615/2020|EPI_ISL_610174|20 ···················· ···················· ····················· ···················· ···················· ····················
N 238 hCoV-19/Hong Kong/HKPU-0442/2020|EPI_ISL_610175|20 ···················· ···················· ····················· ···················· ···················· ····················
N 239 hCoV-19/Hong Kong/HKPU-0613/2020|EPI_ISL_610177|20 ···················· ···················· ····················· ···················· ···················· ····················
N 240 hCoV-19/Hong Kong/HKPU-1362/2020|EPI_ISL_610179|20 ···················· ···················· ····················· ···················· ···················· ····················
N 241 hCoV-19/Hong Kong/HKPU-1326/2020|EPI_ISL_610180|20 ···················· ···················· ····················· ···················· ···················· ····················
N 242 hCoV-19/Hong Kong/HKPU-1319/2020|EPI_ISL_610181|20 ···················· ···················· ····················· ···················· ···················· ····················
N 243 hCoV-19/Hong Kong/HKPU-1448/2020|EPI_ISL_610182|20 ···················· ···················· ····················· ···················· ···················· ····················
N 244 hCoV-19/Hong Kong/HKPU-1470/2020|EPI_ISL_610183|20 ···················· ···················· ····················· ···················· ···················· ····················
N 245 hCoV-19/Hong Kong/HKPU-1570/2020|EPI_ISL_610184|20 ···················· ···················· ····················· ···················· ···················· ····················
N 246 hCoV-19/Hong Kong/HKPU-1296/2020|EPI_ISL_610185|20 ···················· ···················· ····················· ···················· ···················· ····················
N 247 hCoV-19/Hong Kong/HKPU-1366/2020|EPI_ISL_610186|20 ···················· ···················· ····················· ···················· ···················· ····················
N 248 hCoV-19/Hong Kong/HKPU-1402/2020|EPI_ISL_610187|20 ···················· ···················· ····················· ···················· ···················· ····················
N 249 hCoV-19/Hong Kong/HKPU-1428/2020|EPI_ISL_610188|20 ···················· ···················· ····················· ···················· ···················· ····················
N 250 hCoV-19/Hong Kong/HKPU-1437/2020|EPI_ISL_610189|20 ···················· ···················· ····················· ···················· ···················· ····················
N 251 hCoV-19/Hong Kong/HKPU-1540/2020|EPI_ISL_610190|20 ···················· ···················· ····················· ···················· ···················· ····················
N 252 hCoV-19/Hong Kong/HKPU-1465/2020|EPI_ISL_610191|20 ···················· ···················· ····················· ···················· ···················· ····················
N 253 hCoV-19/Hong Kong/HKPU-1433/2020|EPI_ISL_610192|20 ···················· ···················· ····················· ···················· ···················· ····················
N 254 hCoV-19/Hong Kong/HKPU-5052/2020|EPI_ISL_610194|20 ···················· ···················· ····················· ···················· ···················· ····················
N 255 hCoV-19/Hong Kong/HKPU-5073/2020|EPI_ISL_610198|20 ···················· ···················· ····················· ···················· ···················· ····················
N 257 hCoV-19/Ireland/D-NVRL-73IRL84977/2020|EPI_ISL_596 ···················· ···················· ····················· ···················· ···················· ····················
N 258 hCoV-19/Ireland/D-NVRL-73IRL76184/2020|EPI_ISL_596 ···················· ···················· ····················· ···················· ···················· ····················
N 259 hCoV-19/Ireland/RN-NVRL-73IRL88204/2020|EPI_ISL_59 ···················· ···················· ····················· ···················· ···················· ····················
N 260 hCoV-19/Ireland/D-NVRL-73IRL92397/2020|EPI_ISL_596 ···················· ···················· ····················· ···················· ···················· ····················
N 261 hCoV-19/Ireland/D-NVRL-73IRL76176/2020|EPI_ISL_596 ···················· ···················· ····················· ···················· ···················· ····················
N 262 hCoV-19/Ireland/D-NVRL-73IRL76188/2020|EPI_ISL_596 ···················· ···················· ····················· ···················· ···················· ····················
N 263 hCoV-19/Ireland/OY-NVRL-73IRL87287/2020|EPI_ISL_59 ···················· ···················· ····················· ···················· ···················· ····················
N 264 hCoV-19/England/CAMC-A5802D/2020|EPI_ISL_596928|20 ···················· ···················· ····················· ···················· ···················· ····················
N 265 hCoV-19/England/CAMC-A58DF9/2020|EPI_ISL_596929|20 ···················· ···················· ····················· ···················· ···················· ····················
N 266 hCoV-19/England/CAMC-A5844F/2020|EPI_ISL_596932|20 ···················· ·A·················· ····················· ···················· ···················· ···A················
N 267 hCoV-19/England/CAMC-A5945D/2020|EPI_ISL_596933|20 ···················· ···················· ····················· ···················· ···················· ····················
N 268 hCoV-19/England/CAMC-A58EF6/2020|EPI_ISL_596934|20 ···················· ·A·················· ····················· ···················· ···················· ···A················
N 269 hCoV-19/England/CAMC-A58245/2020|EPI_ISL_596935|20 ···················· ···················· ····················· ···················· ···················· ····················
N 270 hCoV-19/England/ALDP-A515E9/2020|EPI_ISL_598000|20 ···················· ···················· ····················· ···················· ···················· ····················
N 271 hCoV-19/England/ALDP-A523FD/2020|EPI_ISL_598002|20 ···················· ···················· ····················· ···················· ···················· ····················
N 272 hCoV-19/England/ALDP-A51F63/2020|EPI_ISL_598003|20 ···················· ···················· ····················· ···················· ······T············· ····················
N 273 hCoV-19/England/ALDP-A51A62/2020|EPI_ISL_598004|20 ···················· ···················· ····················· ···················· ···················· ····················
N 274 hCoV-19/England/ALDP-A51FBE/2020|EPI_ISL_598005|20 ···················· ···················· ····················· ···················· ···················· ····················
N 275 hCoV-19/England/QEUH-A0B180/2020|EPI_ISL_599260|20 ···················· ···················· ····················· ···················· ···················· ····················
N 276 hCoV-19/Scotland/QEUH-A0B609/2020|EPI_ISL_599261|2 ···················· ···················· ····················· ···················· ······T············· ····················
N 277 hCoV-19/England/QEUH-A0C91E/2020|EPI_ISL_599262|20 ···················· ···················· ····················· ···················· ···················· ····················
N 278 hCoV-19/England/QEUH-A0AF87/2020|EPI_ISL_599263|20 ···················· ···················· ····················· ···················· ···················· ····················
N 279 hCoV-19/England/QEUH-A0BA76/2020|EPI_ISL_599264|20 ···················· ···················· ····················· ···················· ···················· ····················
N 280 hCoV-19/England/QEUH-A0C27D/2020|EPI_ISL_599266|20 ···················· ···················· ····················· ···················· ···················· ····················
N 281 hCoV-19/Scotland/QEUH-A0B87C/2020|EPI_ISL_599267|2 ···················· ···················· ····················· ···················· ···················· ····················
N 282 hCoV-19/Scotland/QEUH-A0BD22/2020|EPI_ISL_599268|2 ···················· ···················· ····················· ···················· ···················· ····················
N 283 hCoV-19/England/QEUH-A0C2F5/2020|EPI_ISL_599269|20 ···················· ···················· ····················· ···················· ···················· ····················
N 284 hCoV-19/England/QEUH-A0B9F1/2020|EPI_ISL_599271|20 ···················· ···················· ····················· ···················· ···················· ····················
N 285 hCoV-19/England/QEUH-A0B2BA/2020|EPI_ISL_599272|20 ···················· ···················· ····················· ···················· ···················· ····················
N 286 hCoV-19/England/QEUH-A0B9C4/2020|EPI_ISL_599273|20 ···················· ····A··············· ····················· ···················· ···················· ····················
N 287 hCoV-19/England/QEUH-A0AF2D/2020|EPI_ISL_599275|20 ···················· ···················· ····················· ···················· ···················· ····················
N 288 hCoV-19/Scotland/QEUH-A0C2C8/2020|EPI_ISL_599276|2 ···················· ···················· ····················· ···················· ···················· ····················
N 289 hCoV-19/England/QEUH-A0AD32/2020|EPI_ISL_599277|20 ···················· ···················· ····················· ···················· ···················· ····················
N 290 hCoV-19/England/QEUH-A0AAD1/2020|EPI_ISL_599278|20 ···················· ···················· ····················· ···················· ······T············· ····················
N 291 hCoV-19/Scotland/QEUH-A0B320/2020|EPI_ISL_599279|2 ···················· ···················· ····················· ···················· ···················· ····················
```

| | | | | | | | |
|---|---|---|---|---|---|---|---|
| N | 292 hCoV-19/England/QEUH-A0B0ED/2020\|EPI_ISL_599300\|20 | ···················· | ···················· | ···················· | ···················· | ···················· | ···················· |
| N | 293 hCoV-19/England/QEUH-A0B223/2020\|EPI_ISL_599301\|20 | ···················· | ···················· | ···················· | ···················· | ···················· | ···················· |
| N | 294 hCoV-19/England/QEUH-A0AC35/2020\|EPI_ISL_599302\|20 | ···················· | ·A·················· | ···················· | ···················· | ···················· | ···A················ |
| N | 295 hCoV-19/England/QEUH-A0AB29/2020\|EPI_ISL_599303\|20 | ···················· | ···················· | ···················· | ···················· | ···················· | ···················· |
| N | 296 hCoV-19/England/QEUH-A0B51B/2020\|EPI_ISL_599304\|20 | ···················· | ···················· | ···················· | ···················· | ···················· | ···················· |
| N | 297 hCoV-19/England/QEUH-A0AE7B/2020\|EPI_ISL_599305\|20 | ···················· | ···················· | ···················· | ···················· | ···················· | ···················· |
| N | 298 hCoV-19/Scotland/QEUH-A0B7AC/2020\|EPI_ISL_599306\|2 | ···················· | ···················· | ···················· | ···················· | ···················· | ···················· |
| N | 299 hCoV-19/England/QEUH-A0B8A9/2020\|EPI_ISL_599307\|20 | ···················· | ···················· | ···················· | ···················· | ···················· | ···················· |
| N | 300 hCoV-19/England/QEUH-A0AB74/2020\|EPI_ISL_599308\|20 | ···················· | ···················· | ···················· | ···················· | ···················· | ···················· |
| N | 301 hCoV-19/England/QEUH-A0B733/2020\|EPI_ISL_599309\|20 | ···················· | ···················· | ···················· | ···················· | ···················· | ···················· |
| N | 302 hCoV-19/Scotland/QEUH-A0B2C9/2020\|EPI_ISL_599310\|2 | ···················· | ···················· | ···················· | ···················· | ···················· | ···················· |
| N | 303 hCoV-19/Scotland/QEUH-A0BCE9/2020\|EPI_ISL_599311\|2 | ···················· | ···················· | ···················· | ···················· | ···················· | ···················· |
| N | 304 hCoV-19/England/QEUH-A0B672/2020\|EPI_ISL_599312\|20 | ···················· | ···················· | ···················· | ···················· | ···················· | ···················· |
| N | 305 hCoV-19/England/QEUH-A0B065/2020\|EPI_ISL_599313\|20 | ···················· | ···················· | ···················· | ···················· | ···················· | ···················· |
| N | 306 hCoV-19/England/QEUH-A0B0A1/2020\|EPI_ISL_599315\|20 | ···················· | ···················· | ···················· | ···················· | ···················· | ···················· |
| N | 307 hCoV-19/Scotland/QEUH-A0C2B9/2020\|EPI_ISL_599316\|2 | ···················· | ···················· | ···················· | ···················· | ···················· | ···················· |
| N | 308 hCoV-19/England/QEUH-A0BA67/2020\|EPI_ISL_599317\|20 | ···················· | ···················· | ···················· | ···················· | ···················· | ···················· |
| N | 309 hCoV-19/England/QEUH-A0AD23/2020\|EPI_ISL_599318\|20 | ···················· | ···················· | ···················· | ···················· | ···················· | ···················· |
| N | 310 hCoV-19/England/QEUH-A0B0B0/2020\|EPI_ISL_599319\|20 | ···················· | ···················· | ···················· | ···················· | ···················· | ···················· |
| N | 311 hCoV-19/England/QEUH-A0B135/2020\|EPI_ISL_599322\|20 | ···················· | ···················· | ···················· | ···················· | ···················· | ···················· |
| N | 312 hCoV-19/England/QEUH-A0C899/2020\|EPI_ISL_599323\|20 | ···················· | ···················· | ···················· | ···················· | ······T············· | ···················· |
| N | 313 hCoV-19/England/QEUH-A0B681/2020\|EPI_ISL_599324\|20 | ···················· | ···················· | ···················· | ···················· | ···················· | ···················· |
| N | 314 hCoV-19/Scotland/QEUH-A0BF2C/2020\|EPI_ISL_599325\|2 | ···················· | ···················· | ···················· | ···················· | ···················· | ···················· |
| N | 315 hCoV-19/Scotland/QEUH-A0B92E/2020\|EPI_ISL_599326\|2 | ···················· | ···················· | ···················· | ···················· | ···················· | ···················· |
| N | 316 hCoV-19/England/QEUH-A0AB56/2020\|EPI_ISL_599327\|20 | ···················· | ···················· | ···················· | ···················· | ···················· | ···················· |
| N | 317 hCoV-19/England/QEUH-A0BA85/2020\|EPI_ISL_599328\|20 | ···················· | ···················· | ···················· | ···················· | ···················· | ···················· |
| N | 318 hCoV-19/Wales/QEUH-A0B117/2020\|EPI_ISL_599329\|2020 | ···················· | ···················· | ···················· | ···················· | ···················· | ···················· |
| N | 319 hCoV-19/England/QEUH-A0BF68/2020\|EPI_ISL_599330\|20 | ···················· | ···················· | ···················· | ···················· | ····T··············· | ···················· |
| N | 320 hCoV-19/Scotland/QEUH-A0C2AA/2020\|EPI_ISL_599331\|2 | ···················· | ···················· | ···················· | ···················· | ···················· | ···················· |
| N | 321 hCoV-19/Scotland/QEUH-A0C107/2020\|EPI_ISL_599332\|2 | ···················· | ···················· | ···················· | ···················· | ···················· | ···················· |
| N | 322 hCoV-19/Scotland/QEUH-A0C2E6/2020\|EPI_ISL_599333\|2 | ···················· | ···················· | ···················· | ···················· | ···················· | ···················· |
| N | 323 hCoV-19/England/QEUH-A0B108/2020\|EPI_ISL_599334\|20 | ···················· | ···················· | ···················· | ···················· | ···················· | ···················· |
| N | 324 hCoV-19/England/QEUH-A0B988/2020\|EPI_ISL_599335\|20 | ···················· | ···················· | ···················· | ···················· | ···················· | ···················· |
| N | 325 hCoV-19/England/QEUH-A0AC62/2020\|EPI_ISL_599336\|20 | ···················· | ···················· | ···················· | ···················· | ···················· | ···················· |
| N | 326 hCoV-19/Scotland/QEUH-9F6AD9/2020\|EPI_ISL_600700\|2 | ···················· | ···················· | ···················· | ···················· | ···················· | ···················· |
| N | 327 hCoV-19/Scotland/QEUH-9F4437/2020\|EPI_ISL_600701\|2 | ···················· | ···················· | ···················· | ···················· | ···················· | ···················· |
| N | 328 hCoV-19/Scotland/QEUH-9F44EC/2020\|EPI_ISL_600702\|2 | ···················· | ···················· | ···················· | ···················· | ···················· | ···················· |
| N | 329 hCoV-19/Scotland/QEUH-9F6B12/2020\|EPI_ISL_600704\|2 | ···················· | ···················· | ···················· | ···················· | ···················· | ···················· |
| N | 330 hCoV-19/Scotland/QEUH-9F4A35/2020\|EPI_ISL_600705\|2 | ···················· | ···················· | ···················· | ···················· | ···················· | ···················· |
| N | 331 hCoV-19/Scotland/QEUH-9F6A33/2020\|EPI_ISL_600706\|2 | ···················· | ···················· | ···················· | ···················· | ···················· | ···················· |
| N | 332 hCoV-19/Scotland/QEUH-9F4622/2020\|EPI_ISL_600707\|2 | ···················· | ·A·················· | ···················· | ···················· | ···················· | ···················· |
| N | 333 hCoV-19/England/QEUH-9F477A/2020\|EPI_ISL_600708\|20 | ···················· | ···················· | ···················· | ···················· | ···················· | ···················· |
| N | 334 hCoV-19/Scotland/QEUH-9F5DC2/2020\|EPI_ISL_600709\|2 | ···················· | ···················· | ···················· | ···················· | ···················· | ···················· |
| N | 335 hCoV-19/Scotland/QEUH-9F46B9/2020\|EPI_ISL_600710\|2 | ···················· | ···················· | ···················· | ···················· | ···················· | ···················· |
| N | 336 hCoV-19/Scotland/QEUH-9F4A62/2020\|EPI_ISL_600711\|2 | ···················· | ···················· | ···················· | ···················· | ···················· | ···················· |
| N | 337 hCoV-19/Scotland/QEUH-9F5DFF/2020\|EPI_ISL_600712\|2 | ···················· | ···················· | ···················· | ···················· | ···················· | ···················· |
| N | 338 hCoV-19/Scotland/QEUH-9F681B/2020\|EPI_ISL_600714\|2 | ···················· | ···················· | ···················· | ···················· | ···················· | ···················· |
| N | 339 hCoV-19/Scotland/QEUH-9F673C/2020\|EPI_ISL_600715\|2 | ···················· | ···················· | ···················· | ···················· | ···················· | ···················· |
| N | 340 hCoV-19/Scotland/QEUH-9F6990/2020\|EPI_ISL_600716\|2 | ···················· | ···················· | ···················· | ···················· | ···················· | ···················· |
| N | 341 hCoV-19/Scotland/QEUH-9F4534/2020\|EPI_ISL_600717\|2 | ···················· | ···················· | ···················· | ···················· | ···················· | ···················· |
| N | 342 hCoV-19/Scotland/QEUH-9F4CD5/2020\|EPI_ISL_600718\|2 | ···················· | ···················· | ···················· | ···················· | ···················· | ···················· |
| N | 343 hCoV-19/Scotland/QEUH-9F468C/2020\|EPI_ISL_600720\|2 | ···················· | ···················· | ···················· | ···················· | ···················· | ···················· |
| N | 344 hCoV-19/Scotland/QEUH-9F5DB3/2020\|EPI_ISL_600721\|2 | ···················· | ···················· | ···················· | ···················· | ···················· | ···················· |
| N | 345 hCoV-19/Scotland/QEUH-9F4CC6/2020\|EPI_ISL_600723\|2 | ···················· | ···················· | ···················· | ···················· | ···················· | ···················· |
| N | 346 hCoV-19/Scotland/QEUH-9F6945/2020\|EPI_ISL_600724\|2 | ···················· | ···················· | ···················· | ···················· | ···················· | ···················· |
| N | 347 hCoV-19/Scotland/QEUH-9F6866/2020\|EPI_ISL_600725\|2 | ···················· | ···················· | ···················· | ···················· | ···················· | ···················· |
| N | 348 hCoV-19/Scotland/QEUH-9F670F/2020\|EPI_ISL_600726\|2 | ···················· | ···················· | ···················· | ···················· | ···················· | ···················· |
| N | 349 hCoV-19/Scotland/QEUH-9F440A/2020\|EPI_ISL_600727\|2 | ···················· | ···················· | ···················· | ···················· | ···················· | ···················· |
| N | 350 hCoV-19/Scotland/QEUH-9F4798/2020\|EPI_ISL_600728\|2 | ···················· | ···················· | ···················· | ···················· | ···················· | ···················· |
| N | 351 hCoV-19/Scotland/QEUH-9F4640/2020\|EPI_ISL_600729\|2 | ···················· | ···················· | ···················· | ···················· | ···················· | ···················· |
| N | 352 hCoV-19/Scotland/QEUH-9F4CA8/2020\|EPI_ISL_600730\|2 | ···················· | ···················· | ···················· | ···················· | ···················· | ···················· |
| N | 353 hCoV-19/England/QEUH-9F44BF/2020\|EPI_ISL_600731\|20 | ···················· | ···················· | ···················· | ···················· | ···················· | ···················· |
| N | 354 hCoV-19/Scotland/QEUH-9F6839/2020\|EPI_ISL_600732\|2 | ···················· | ···················· | ···················· | ···················· | ···················· | ···················· |
| N | 355 hCoV-19/Scotland/QEUH-9F4604/2020\|EPI_ISL_600733\|2 | ···················· | ···················· | ···················· | ···················· | ···················· | ···················· |
| N | 356 hCoV-19/Scotland/QEUH-9F4CB7/2020\|EPI_ISL_600734\|2 | ···················· | ···················· | ···················· | ···················· | ···················· | ···················· |
| N | 357 hCoV-19/Scotland/QEUH-9F469B/2020\|EPI_ISL_600735\|2 | ···················· | ···················· | ···················· | ···················· | ···················· | ···················· |
| N | 358 hCoV-19/Scotland/QEUH-9F66E4/2020\|EPI_ISL_600736\|2 | ···················· | ···················· | ···················· | ···················· | ···················· | ···················· |
| N | 359 hCoV-19/Scotland/QEUH-9F6848/2020\|EPI_ISL_600737\|2 | ···················· | ···················· | ···················· | ···················· | ···················· | ···················· |
| N | 360 hCoV-19/Scotland/QEUH-9F4CE4/2020\|EPI_ISL_600738\|2 | ···················· | ···················· | ···················· | ···················· | ···················· | ···················· |
| N | 361 hCoV-19/Scotland/QEUH-9F4507/2020\|EPI_ISL_600741\|2 | ···················· | ···················· | ···················· | ···················· | ···················· | ···················· |
| N | 362 hCoV-19/Scotland/QEUH-9F45E9/2020\|EPI_ISL_600742\|2 | ···················· | ···················· | ···················· | ···················· | ···················· | ···················· |
| N | 363 hCoV-19/Scotland/QEUH-9F6B03/2020\|EPI_ISL_600743\|2 | ···················· | ···················· | ···················· | ···················· | ···················· | ···················· |
| N | 364 hCoV-19/Scotland/QEUH-9F6769/2020\|EPI_ISL_600744\|2 | ···················· | ···················· | ···················· | ···················· | ···················· | ···················· |
| N | 365 hCoV-19/Scotland/QEUH-9F6A8E/2020\|EPI_ISL_600745\|2 | ···················· | ···················· | ···················· | ···················· | ···················· | ···················· |
| N | 366 hCoV-19/Scotland/QEUH-9F4631/2020\|EPI_ISL_600746\|2 | ···················· | ···················· | ···················· | ···················· | ···················· | ···················· |
| N | 367 hCoV-19/Scotland/QEUH-9F4A44/2020\|EPI_ISL_600747\|2 | ···················· | ···················· | ···················· | ···················· | ···················· | ···················· |
| N | 368 hCoV-19/Scotland/QEUH-9F65F6/2020\|EPI_ISL_600748\|2 | ···················· | ···················· | ···················· | ···················· | ···················· | ···················· |
| N | 369 hCoV-19/Scotland/QEUH-9F46AA/2020\|EPI_ISL_600749\|2 | ···················· | ···················· | ···················· | ···················· | ···················· | ···················· |
| N | 370 hCoV-19/Scotland/QEUH-9F672D/2020\|EPI_ISL_600750\|2 | ···················· | ···················· | ···················· | ···················· | ···················· | ···················· |
| N | 371 hCoV-19/Scotland/QEUH-9F4543/2020\|EPI_ISL_600751\|2 | ···················· | ···················· | ···················· | ···················· | ···················· | ···················· |

```
N    372 hCoV-19/Scotland/QEUH-9F6AF7/2020|EPI_ISL_600752|2    ····················  ····················  ·····················  ····················  ····················  ····················
N    373 hCoV-19/Scotland/QEUH-9F69CD/2020|EPI_ISL_600753|2    ····················  ····················  ·····················  ····················  ····················  ····················
N    374 hCoV-19/Scotland/QEUH-9F5E47/2020|EPI_ISL_600754|2    ····················  ····················  ·····················  ····················  ····················  ····················
N    375 hCoV-19/Scotland/QEUH-9F67D2/2020|EPI_ISL_600755|2    ····················  ····················  ·····················  ····················  ····················  ····················
N    376 hCoV-19/Scotland/QEUH-9F5F62/2020|EPI_ISL_600756|2    ····················  ····················  ·····················  ····················  ····················  ····················
N    377 hCoV-19/Scotland/QEUH-9F5D68/2020|EPI_ISL_600757|2    ····················  ····················  ·····················  ····················  ····················  ····················
N    378 hCoV-19/Scotland/QEUH-9F6936/2020|EPI_ISL_600758|2    ····················  ····················  ·····················  ····················  ····················  ····················
N    379 hCoV-19/Scotland/QEUH-9F2D10/2020|EPI_ISL_600760|2    ····················  ····················  ·····················  ····················  ······T·············  ····················
N    380 hCoV-19/Scotland/QEUH-9F42B5/2020|EPI_ISL_600761|2    ····················  ····················  ·····················  ····················  ····················  ····················
N    381 hCoV-19/Scotland/QEUH-9F3ABE/2020|EPI_ISL_600762|2    ····················  ····················  ·····················  ····················  ····················  ····················
N    382 hCoV-19/Scotland/QEUH-9F309E/2020|EPI_ISL_600763|2    ····················  ····················  ·····················  ····················  ····················  ····················
N    383 hCoV-19/Scotland/QEUH-9F3799/2020|EPI_ISL_600764|2    ····················  ······C·············  ·····················  ····················  ····················  ····················
N    384 hCoV-19/Scotland/QEUH-9F3A72/2020|EPI_ISL_600765|2    ····················  ····················  ·····················  ····················  ····················  ····················
N    385 hCoV-19/Scotland/QEUH-9F37D5/2020|EPI_ISL_600766|2    ····················  ····················  ·····················  ····················  ····················  ····················
N    386 hCoV-19/Scotland/QEUH-9F3DE2/2020|EPI_ISL_600767|2    ····················  ····················  ·····················  ····················  ····················  ····················
N    387 hCoV-19/Scotland/QEUH-9F2D5C/2020|EPI_ISL_600768|2    ····················  ····················  ·····················  ····················  ····················  ····················
N    388 hCoV-19/England/QEUH-9F2C31/2020|EPI_ISL_600769|20    ····················  ····················  ·····················  ····················  ····················  ····················
N    389 hCoV-19/England/QEUH-9F38F0/2020|EPI_ISL_600770|20    ····················  ····················  ·····················  ····················  ····················  ····················
N    390 hCoV-19/Scotland/QEUH-9F3113/2020|EPI_ISL_600771|2    ····················  ····················  ·····················  ····················  ····················  ····················
N    391 hCoV-19/England/QEUH-9F3034/2020|EPI_ISL_600772|20    ····················  ····················  ·····················  ····················  ····················  ····················
N    392 hCoV-19/Scotland/QEUH-9F2D01/2020|EPI_ISL_600773|2    ····················  ····················  ·····················  ····················  ····················  ····················
N    393 hCoV-19/Scotland/QEUH-9F3131/2020|EPI_ISL_600774|2    ····················  ····················  ·····················  ····················  ····················  ····················
N    394 hCoV-19/England/QEUH-9F3043/2020|EPI_ISL_600775|20    ····················  ····················  ·····················  ····················  ····················  ····················
N    395 hCoV-19/Scotland/QEUH-9F30BC/2020|EPI_ISL_600776|2    ····················  ····················  ·····················  ····················  ····················  ····················
N    396 hCoV-19/Scotland/QEUH-9F3FCE/2020|EPI_ISL_600777|2    ····················  ····················  ·····················  ····················  ····················  ····················
N    397 hCoV-19/England/QEUH-9F3B7F/2020|EPI_ISL_600778|20    ····················  ·A··················  ·····················  ····················  ····················  ···A················
N    398 hCoV-19/England/QEUH-9F3289/2020|EPI_ISL_600779|20    ····················  ·A··················  ·····················  ····················  ····················  ···A················
N    399 hCoV-19/Scotland/QEUH-9F2CE6/2020|EPI_ISL_600780|2    ····················  ····················  ·····················  ····················  ····················  ····················
N    400 hCoV-19/Scotland/QEUH-9F32E3/2020|EPI_ISL_600781|2    ····················  ····················  ·····················  ····················  ····················  ····················
N    401 hCoV-19/England/QEUH-9F39DF/2020|EPI_ISL_600782|20    ····················  ····················  ·····················  ····················  ····················  ····················
N    402 hCoV-19/England/QEUH-9F4288/2020|EPI_ISL_600783|20    ····················  ····················  ·····················  ····················  ····················  ····················
N    403 hCoV-19/England/QEUH-9F3CE5/2020|EPI_ISL_600785|20    ····················  ····················  ·····················  ····················  ····················  ····················
N    404 hCoV-19/England/QEUH-9F3052/2020|EPI_ISL_600786|20    ····················  ····················  ·····················  ····················  ····················  ····················
N    405 hCoV-19/Scotland/QEUH-9F3D00/2020|EPI_ISL_600787|2    ····················  ····················  ·····················  ····················  ····················  ····················
N    406 hCoV-19/England/QEUH-9F426A/2020|EPI_ISL_600788|20    ····················  ····················  ·····················  ····················  ····················  ····················
N    407 hCoV-19/Scotland/QEUH-9F42C4/2020|EPI_ISL_600789|2    ····················  ····················  ·····················  ····················  ····················  ····················
N    408 hCoV-19/Scotland/QEUH-9F42A6/2020|EPI_ISL_600790|2    ····················  ····················  ·····················  ····················  ····················  ····················
N    409 hCoV-19/Scotland/QEUH-9F2CF5/2020|EPI_ISL_600791|2    ····················  ····················  ·····················  ····················  ····················  ····················
N    410 hCoV-19/Scotland/QEUH-9F3D6A/2020|EPI_ISL_600792|2    ····················  ····················  ·····················  ····················  ····················  ····················
N    411 hCoV-19/Scotland/QEUH-9F3ED0/2020|EPI_ISL_600793|2    ····················  ····················  ·····················  ····················  ····················  ····················
N    412 hCoV-19/Scotland/QEUH-9F2CAA/2020|EPI_ISL_600794|2    ····················  ····················  ·····················  ····················  ····················  ····················
N    413 hCoV-19/England/QEUH-9F2C7D/2020|EPI_ISL_600795|20    ····················  ····················  ·····················  ····················  ····················  ····················
N    414 hCoV-19/Scotland/QEUH-9F3D1F/2020|EPI_ISL_600796|2    ····················  ····················  ·····················  ····················  ····················  ····················
N    415 hCoV-19/Scotland/QEUH-9F4394/2020|EPI_ISL_600797|2    ····················  ····················  ·····················  ····················  ····················  ····················
N    416 hCoV-19/Wales/QEUH-9F37C6/2020|EPI_ISL_600798|2020    ····················  ····················  ·····················  ····················  ····················  ····················
N    417 hCoV-19/Scotland/QEUH-9F3122/2020|EPI_ISL_600799|2    ····················  ····················  ·····················  ····················  ····················  ····················
N    418 hCoV-19/USA/MN-MDH-1865/2020|EPI_ISL_603000|2020-0    ····················  ····················  ·····················  ····················  ····················  ····················
N    419 hCoV-19/USA/MN-MDH-1866/2020|EPI_ISL_603001|2020-0    ····················  ····················  ·····················  ····················  ····················  ····················
N    420 hCoV-19/USA/MN-MDH-1871/2020|EPI_ISL_603006|2020-1    ····················  ·A··················  ·····················  ····················  ····················  ····················
N    421 hCoV-19/USA/MN-MDH-1875/2020|EPI_ISL_603010|2020-0    ····················  ····················  ·····················  ····················  ····················  ····················
N    422 hCoV-19/USA/MN-MDH-1876/2020|EPI_ISL_603011|2020-0    ····················  ····················  ·····················  ····················  ····················  ····················
N    423 hCoV-19/USA/MN-MDH-1877/2020|EPI_ISL_603012|2020-1    ····················  ·A··················  ·····················  ····················  ····················  ····················
N    424 hCoV-19/USA/MN-MDH-1878/2020|EPI_ISL_603013|2020-1    ····················  ·A··················  ·····················  ····················  ····················  ····················
N    425 hCoV-19/USA/MN-MDH-1879/2020|EPI_ISL_603014|2020-1    ····················  ·A··················  ·····················  ····················  ····················  ····················
N    426 hCoV-19/USA/MN-MDH-1880/2020|EPI_ISL_603015|2020-1    ····················  ····················  ·····················  ····················  ····················  ····················
N    427 hCoV-19/USA/MN-MDH-1881/2020|EPI_ISL_603016|2020-1    ····················  ·A··················  ·····················  ····················  ····················  ····················
N    428 hCoV-19/USA/MN-MDH-1882/2020|EPI_ISL_603017|2020-1    ····················  ·A··················  ·····················  ····················  ····················  ····················
N    429 hCoV-19/USA/MN-MDH-1883/2020|EPI_ISL_603018|2020-1    ····················  ····················  ·····················  ····················  ····················  ····················
N    430 hCoV-19/USA/MN-MDH-1884/2020|EPI_ISL_603019|2020-1    ····················  ····················  ·····················  ····················  ··NN················  ····················
N    431 hCoV-19/USA/MN-MDH-1885/2020|EPI_ISL_603020|2020-1    ····················  ····················  ·····················  ····················  ····················  ····················
N    432 hCoV-19/Brazil/SP-427/2020|EPI_ISL_603021|2020-07-    ····················  ····················  ·····················  ····················  ····················  ····················
N    433 hCoV-19/Brazil/SP-429/2020|EPI_ISL_603022|2020-07-    ····················  ····················  ·····················  ····················  ····················  ····················
N    434 hCoV-19/Brazil/SP-376/2020|EPI_ISL_603023|2020-06-    ····················  ····················  ·····················  ····················  ····················  ····················
N    435 hCoV-19/Brazil/SP-383/2020|EPI_ISL_603024|2020-06-    ····················  ····················  ·····················  ····················  ····················  ····················
N    436 hCoV-19/Brazil/SP-384/2020|EPI_ISL_603025|2020-06-    ····················  ····················  ·····················  ····················  ····················  ····················
N    437 hCoV-19/Brazil/SP-385/2020|EPI_ISL_603026|2020-06-    ····················  ····················  ·····················  ····················  ····················  ····················
N    438 hCoV-19/Brazil/SP-387/2020|EPI_ISL_603027|2020-06-    ····················  ····················  ·····················  ····················  ····················  ····················
N    439 hCoV-19/Brazil/SP-398/2020|EPI_ISL_603028|2020-06-    ····················  ····················  ·····················  ····················  ····················  ····················
N    440 hCoV-19/Brazil/SP-399/2020|EPI_ISL_603029|2020-06-    ····················  ····················  ·····················  ····················  ····················  ····················
N    441 hCoV-19/USA/PA-QDX-1951/2020|EPI_ISL_604280|2020-0    ····················  ····················  ·····················  ····················  ····················  ····················
N    442 hCoV-19/USA/UT-QDX-1880/2020|EPI_ISL_604281|2020-0    ····················  ····················  ·····················  ····················  ····················  ····················
N    443 hCoV-19/USA/UT-QDX-1896/2020|EPI_ISL_604282|2020-0    ····················  ····················  ·····················  ····················  ····················  ····················
N    444 hCoV-19/USA/UT-QDX-1901/2020|EPI_ISL_604283|2020-0    ····················  ····················  ·····················  ····················  ····················  ····················
N    445 hCoV-19/USA/UT-QDX-1906/2020|EPI_ISL_604284|2020-0    ····················  ····················  ·····················  ····················  ····················  ····················
N    446 hCoV-19/USA/UT-QDX-1930/2020|EPI_ISL_604285|2020-0    ····················  ····················  ·····················  ····················  ····················  ····················
N    447 hCoV-19/USA/UT-QDX-1936/2020|EPI_ISL_604286|2020-0    ····················  ····················  ·····················  ····················  ····················  ····················
N    448 hCoV-19/USA/UT-QDX-1950/2020|EPI_ISL_604287|2020-0    ····················  ····················  ·····················  ····················  ····················  ····················
N    449 hCoV-19/USA/UT-QDX-1954/2020|EPI_ISL_604288|2020-0    ····················  ····················  ·····················  ····················  ····················  ····················
N    450 hCoV-19/USA/UT-QDX-1959/2020|EPI_ISL_604289|2020-0    ····················  ····················  ·····················  ····················  ····················  ····················
N    451 hCoV-19/USA/UT-QDX-1964/2020|EPI_ISL_604290|2020-0    ····················  ····················  ·····················  ····················  ····················  ····················
```

| | | | | | | | |
|---|---|---|---|---|---|---|---|
| N | 452 hCoV-19/USA/UT-QDX-1966/2020\|EPI_ISL_604291\|2020-0 | ···················· | ···················· | ···················· | ···················· | ···················· | ···················· |
| N | 453 hCoV-19/USA/UT-QDX-1971/2020\|EPI_ISL_604292\|2020-0 | ···················· | ···················· | ···················· | ···················· | ···················· | ···················· |
| N | 454 hCoV-19/USA/CA-QDX-1980/2020\|EPI_ISL_604293\|2020-0 | ···················· | ···················· | ···················· | ···················· | ···················· | ···················· |
| N | 455 hCoV-19/USA/CA-QDX-1981/2020\|EPI_ISL_604294\|2020-0 | ···················· | ···················· | ···················· | ···················· | ···················· | ···················· |
| N | 456 hCoV-19/USA/CA-QDX-1982/2020\|EPI_ISL_604295\|2020-0 | ···················· | ···················· | ···················· | ···················· | ···················· | ···················· |
| N | 457 hCoV-19/USA/CA-QDX-1984/2020\|EPI_ISL_604296\|2020-0 | ···················· | ···················· | ···················· | ···················· | ···················· | ···················· |
| N | 458 hCoV-19/USA/CA-QDX-1986/2020\|EPI_ISL_604297\|2020-0 | ···················· | ···················· | ···················· | ···················· | ···················· | ···················· |
| N | 459 hCoV-19/USA/CA-QDX-1994/2020\|EPI_ISL_604298\|2020-0 | ···················· | ···················· | ···················· | ···················· | ···················· | ···················· |
| N | 460 hCoV-19/USA/CA-QDX-1995/2020\|EPI_ISL_604299\|2020-0 | ···················· | ···················· | ···················· | ···················· | ···················· | ···················· |
| N | 461 hCoV-19/USA/ID-QDX-2061/2020\|EPI_ISL_604330\|2020-0 | ···················· | ···················· | ···················· | ···················· | ···················· | ···················· |
| N | 462 hCoV-19/USA/IN-QDX-1987/2020\|EPI_ISL_604331\|2020-0 | ···················· | ···················· | ···················· | ···················· | ···················· | ···················· |
| N | 463 hCoV-19/USA/IN-QDX-1990/2020\|EPI_ISL_604332\|2020-0 | ···················· | ···················· | ···················· | ···················· | ···················· | ···················· |
| N | 464 hCoV-19/USA/IN-QDX-2023/2020\|EPI_ISL_604333\|2020-0 | ···················· | ···················· | ···················· | ···················· | ···················· | ···················· |
| N | 465 hCoV-19/USA/LA-QDX-1998/2020\|EPI_ISL_604337\|2020-0 | ···················· | ···················· | ···················· | ···················· | ···················· | ···················· |
| N | 466 hCoV-19/USA/LA-QDX-2055/2020\|EPI_ISL_604340\|2020-0 | ···················· | ···················· | ···················· | ···················· | ···················· | ···················· |
| N | 467 hCoV-19/USA/LA-QDX-2056/2020\|EPI_ISL_604341\|2020-0 | ···················· | ···················· | ···················· | ···················· | ···················· | ···················· |
| N | 468 hCoV-19/USA/LA-QDX-2062/2020\|EPI_ISL_604342\|2020-0 | ···················· | ···················· | ···················· | ···················· | ···················· | ···················· |
| N | 469 hCoV-19/USA/MA-QDX-1988/2020\|EPI_ISL_604343\|2020-0 | ···················· | ···················· | ···················· | ···················· | ···················· | ···················· |
| N | 470 hCoV-19/USA/MA-QDX-1991/2020\|EPI_ISL_604344\|2020-0 | ···················· | ···················· | ···················· | ···················· | ···················· | ···················· |
| N | 471 hCoV-19/USA/MA-QDX-2015/2020\|EPI_ISL_604345\|2020-0 | ···················· | ···················· | ···················· | ···················· | ···················· | ···················· |
| N | 472 hCoV-19/USA/MA-QDX-2064/2020\|EPI_ISL_604346\|2020-0 | ···················· | ···················· | ···················· | ···················· | ···················· | ···················· |
| N | 473 hCoV-19/USA/MN-QDX-1996/2020\|EPI_ISL_604347\|2020-0 | ···················· | ···················· | ···················· | ···················· | ···················· | ···················· |
| N | 474 hCoV-19/USA/MN-QDX-2011/2020\|EPI_ISL_604348\|2020-0 | ···················· | ···················· | ···················· | ···················· | ···················· | ···················· |
| N | 475 hCoV-19/USA/MN-QDX-2030/2020\|EPI_ISL_604349\|2020-0 | ···················· | ···················· | ···················· | ···················· | ···················· | ···················· |
| N | 476 hCoV-19/USA/MN-QDX-2036/2020\|EPI_ISL_604350\|2020-0 | ···················· | ···················· | ···················· | ···················· | ···················· | ···················· |
| N | 477 hCoV-19/USA/MN-QDX-2040/2020\|EPI_ISL_604351\|2020-0 | ···················· | ···················· | ···················· | ···················· | ···················· | ···················· |
| N | 478 hCoV-19/USA/MN-QDX-2041/2020\|EPI_ISL_604352\|2020-0 | ···················· | ···················· | ···················· | ···················· | ···················· | ···················· |
| N | 479 hCoV-19/USA/MN-QDX-2042/2020\|EPI_ISL_604353\|2020-0 | ···················· | ···················· | ···················· | ···················· | ···················· | ···················· |
| N | 480 hCoV-19/USA/MN-QDX-2053/2020\|EPI_ISL_604354\|2020-0 | ···················· | ···················· | ···················· | ···················· | ···················· | ···················· |
| N | 481 hCoV-19/USA/MN-QDX-2054/2020\|EPI_ISL_604355\|2020-0 | ···················· | ···················· | ···················· | ···················· | ···················· | ···················· |
| N | 482 hCoV-19/USA/MN-QDX-2059/2020\|EPI_ISL_604356\|2020-0 | ···················· | ···················· | ···················· | ···················· | ···················· | ···················· |
| N | 483 hCoV-19/USA/NY-QDX-1983/2020\|EPI_ISL_604357\|2020-0 | ···················· | ···················· | ···················· | ···················· | ···················· | ···················· |
| N | 484 hCoV-19/USA/NY-QDX-1985/2020\|EPI_ISL_604358\|2020-0 | ···················· | ···················· | ···················· | ···················· | ···················· | ···················· |
| N | 485 hCoV-19/USA/NY-QDX-1993/2020\|EPI_ISL_604359\|2020-0 | ···················· | ···················· | ···················· | ···················· | ···················· | ···················· |
| N | 486 hCoV-19/USA/WI-UW-1631/2020\|EPI_ISL_605505\|2020-09 | ···················· | ···················· | ···················· | ···················· | ···················· | ···················· |
| N | 487 hCoV-19/USA/WI-UW-1632/2020\|EPI_ISL_605506\|2020-09 | ···················· | ···················· | ···················· | ···················· | ···················· | ···················· |
| N | 488 hCoV-19/USA/WI-UW-1633/2020\|EPI_ISL_605507\|2020-09 | ···················· | ···················· | ···················· | ···················· | ···················· | ···················· |
| N | 500 hCoV-19/USA/WI-UW-1804/2020\|EPI_ISL_605676\|2020-09 | ···················· | ···················· | ···················· | ···················· | ···················· | ···················· |
| N | 501 hCoV-19/USA/WI-UW-1805/2020\|EPI_ISL_605677\|2020-09 | ···················· | ···················· | ···················· | ···················· | ···················· | ···················· |
| N | 505 hCoV-19/USA/WI-UW-1809/2020\|EPI_ISL_605681\|2020-09 | ···················· | ···················· | ···················· | ···················· | ···················· | ···················· |
| N | 513 hCoV-19/Ireland/D-NVRL-73IRL76165/2020\|EPI_ISL_596 | ···················· | ···················· | ···················· | ···················· | ···················· | ···················· |
| N | 514 hCoV-19/Ireland/D-NVRL-73IRL76166/2020\|EPI_ISL_596 | ···················· | ···················· | ···················· | ···················· | ···················· | ···················· |
| N | 515 hCoV-19/Ireland/D-NVRL-73IRL76171/2020\|EPI_ISL_596 | ···················· | ···················· | ···················· | ···················· | ···················· | ···················· |
| N | 516 hCoV-19/Ireland/D-NVRL-73IRL37614/2020\|EPI_ISL_596 | ···················· | ···················· | ···················· | ···················· | ···················· | ···················· |
| N | 517 hCoV-19/Ireland/D-NVRL-73IRL76167/2020\|EPI_ISL_596 | ···················· | ···················· | ···················· | ···················· | ···················· | ···················· |
| N | 518 hCoV-19/Ireland/TA-NVRL-73IRL43192/2020\|EPI_ISL_59 | ···················· | ···················· | ···················· | ···················· | ···················· | ···················· |
| N | 519 hCoV-19/Ireland/D-NVRL-73IRL37625/2020\|EPI_ISL_596 | ···················· | ···················· | ···················· | ···················· | ···················· | ···················· |
| N | 520 hCoV-19/Ireland/LH-NVRL-73IRL45658/2020\|EPI_ISL_59 | ···················· | ···················· | ···················· | ···················· | ···················· | ···················· |
| N | 521 hCoV-19/Ireland/LH-NVRL-73IRL41505/2020\|EPI_ISL_59 | ···················· | ···················· | ···················· | ···················· | ···················· | ···················· |
| N | 522 hCoV-19/Ireland/D-NVRL-73IRL76207/2020\|EPI_ISL_596 | ···················· | ···················· | ···················· | ···················· | ···················· | ···················· |
| N | 523 hCoV-19/Ireland/KY-NVRL-73IRL83928/2020\|EPI_ISL_59 | ···················· | ···················· | ···················· | ···················· | ···················· | ···················· |
| N | 524 hCoV-19/Ireland/D-NVRL-73IRL76194/2020\|EPI_ISL_596 | ···················· | ···················· | ···················· | ···················· | ···················· | ···················· |
| N | 525 hCoV-19/Ireland/LK-NVRL-73IRL80542/2020\|EPI_ISL_59 | ···················· | ·A·················· | ···················· | ···················· | ···················· | ··A················· |
| N | 526 hCoV-19/Ireland/D-NVRL-73IRL76233/2020\|EPI_ISL_596 | ···················· | ···················· | ···················· | ···················· | ···················· | ···················· |
| N | 527 hCoV-19/Ireland/D-NVRL-73IRL85097/2020\|EPI_ISL_596 | ···················· | ···················· | ···················· | ···················· | ···················· | ···················· |
| N | 528 hCoV-19/Ireland/D-NVRL-73IRL86995/2020\|EPI_ISL_596 | ···················· | ···················· | ···················· | ···················· | ···················· | ···················· |
| N | 529 hCoV-19/Ireland/D-NVRL-73IRL76182/2020\|EPI_ISL_596 | ···················· | ···················· | ···················· | ···················· | ···················· | ···················· |
| N | 530 hCoV-19/Ireland/KE-NVRL-73IRL78912/2020\|EPI_ISL_59 | ···················· | ···················· | ···················· | ···················· | ···················· | ···················· |
| N | 531 hCoV-19/Ireland/D-NVRL-73IRL93118/2020\|EPI_ISL_596 | ···················· | ···················· | ···················· | ···················· | ···················· | ···················· |
| N | 532 hCoV-19/England/CAMC-A588F8/2020\|EPI_ISL_596930\|20 | ···················· | ···················· | ···················· | ···················· | ···················· | ···················· |
| N | 533 hCoV-19/England/CAMC-A57E8E/2020\|EPI_ISL_596931\|20 | ···················· | ···················· | ···················· | ···················· | ···················· | ···················· |
| N | 534 hCoV-19/England/CAMC-A58A7A/2020\|EPI_ISL_596936\|20 | ···················· | ···················· | ···················· | ···················· | ···················· | ···················· |
| N | 535 hCoV-19/England/CAMC-A58193/2020\|EPI_ISL_596938\|20 | ···················· | ···················· | ···················· | ···················· | ···················· | ···················· |
| N | 536 hCoV-19/England/CAMC-A5855B/2020\|EPI_ISL_596940\|20 | ···················· | ···················· | ···················· | ···················· | ···················· | ···················· |
| N | 537 hCoV-19/England/CAMC-A57F03/2020\|EPI_ISL_596941\|20 | ···················· | ···················· | ···················· | ···················· | ···················· | ···················· |
| N | 538 hCoV-19/England/CAMC-A58825/2020\|EPI_ISL_596942\|20 | ···················· | ···················· | ···················· | ···················· | ···················· | ···················· |
| N | 539 hCoV-19/England/CAMC-A580B4/2020\|EPI_ISL_596944\|20 | ···················· | ···················· | ···················· | ···················· | ···················· | ···················· |
| N | 540 hCoV-19/England/CAMC-A5845E/2020\|EPI_ISL_596945\|20 | ···················· | ···················· | ···················· | ···················· | ······T············· | ···················· |
| N | 541 hCoV-19/England/CAMC-A57DCD/2020\|EPI_ISL_596946\|20 | ···················· | ···················· | ···················· | ···················· | ···················· | ···················· |
| N | 542 hCoV-19/England/CAMC-A58D53/2020\|EPI_ISL_596947\|20 | ···················· | ···················· | ···················· | ···················· | ···················· | ···················· |
| N | 543 hCoV-19/England/CAMC-A58940/2020\|EPI_ISL_596951\|20 | ···················· | ···················· | ···················· | ···················· | ···················· | ···················· |
| N | 544 hCoV-19/England/CAMC-A58306/2020\|EPI_ISL_596952\|20 | ···················· | ···················· | ···················· | ···················· | ···················· | ···················· |
| N | 545 hCoV-19/England/CAMC-A5870A/2020\|EPI_ISL_596953\|20 | ···················· | ···················· | ···················· | ···················· | ···················· | ···················· |
| N | 546 hCoV-19/England/CAMC-A5948A/2020\|EPI_ISL_596954\|20 | ···················· | ···················· | ···················· | ···················· | ···················· | ···················· |
| N | 547 hCoV-19/England/CAMC-A5946C/2020\|EPI_ISL_596955\|20 | ···················· | ···················· | ···················· | ···················· | ···················· | ···················· |
| N | 548 hCoV-19/England/CAMC-A585A6/2020\|EPI_ISL_596956\|20 | ···················· | ···················· | ···················· | ···················· | ···················· | ···················· |
| N | 549 hCoV-19/England/CAMC-A591CF/2020\|EPI_ISL_596957\|20 | ···················· | ···················· | ···················· | ···················· | ···················· | ···················· |
| N | 550 hCoV-19/England/CAMC-A57D27/2020\|EPI_ISL_596958\|20 | ···················· | ···················· | ···················· | ···················· | ······T············· | ···················· |
| N | 551 hCoV-19/England/CAMC-A58CCF/2020\|EPI_ISL_596959\|20 | ···················· | ···················· | ···················· | ···················· | ···················· | ···················· |
| N | 552 hCoV-19/England/CAMC-A58D71/2020\|EPI_ISL_596960\|20 | ···················· | ···················· | ···················· | ···················· | ···················· | ···················· |

| | | | | | | | | |
|---|---|---|---|---|---|---|---|---|
| N | 553 | hCoV-19/England/CAMC-A5803C/2020\|EPI_ISL_596961\|20 | ···················· | ···················· | ···················· | ···················· | ···················· | ···················· |
| N | 554 | hCoV-19/England/CAMC-A580A5/2020\|EPI_ISL_596962\|20 | ···················· | ···················· | ···················· | ···················· | ···················· | ···················· |
| N | 555 | hCoV-19/England/CAMC-A58A01/2020\|EPI_ISL_596963\|20 | ···················· | ···················· | ···················· | ···················· | ···················· | ···················· |
| N | 556 | hCoV-19/England/CAMC-A58F99/2020\|EPI_ISL_596964\|20 | ···················· | ···················· | ···················· | ···················· | ···················· | ···················· |
| N | 557 | hCoV-19/England/CAMC-A58649/2020\|EPI_ISL_596965\|20 | ···················· | ···················· | ···················· | ···················· | ···················· | ···················· |
| N | 558 | hCoV-19/England/CAMC-A58B3B/2020\|EPI_ISL_596966\|20 | ···················· | ···················· | ···················· | ···················· | ···················· | ···················· |
| N | 559 | hCoV-19/England/CAMC-A582FA/2020\|EPI_ISL_596967\|20 | ···················· | ···················· | ···················· | ···················· | ···················· | ···················· |
| N | 560 | hCoV-19/England/CAMC-A5962A/2020\|EPI_ISL_596968\|20 | ···················· | ···················· | ···················· | ···················· | ···················· | ···················· |
| N | 561 | hCoV-19/England/CAMC-A596EE/2020\|EPI_ISL_596970\|20 | ···················· | ·A·················· | ···················· | ···················· | ···················· | ··A················· |
| N | 562 | hCoV-19/England/CAMC-A581FD/2020\|EPI_ISL_596971\|20 | ···················· | ···················· | ···················· | ···················· | ···················· | ···················· |
| N | 563 | hCoV-19/England/CAMC-A595D2/2020\|EPI_ISL_596972\|20 | ···················· | ···················· | ···················· | ···················· | ···················· | ···················· |
| N | 564 | hCoV-19/England/CAMC-A57E7F/2020\|EPI_ISL_596974\|20 | ···················· | ···················· | ···················· | ···················· | ···················· | ···················· |
| N | 565 | hCoV-19/England/CAMC-A5847C/2020\|EPI_ISL_596975\|20 | ···················· | ···················· | ···················· | ···················· | ···················· | ···················· |
| N | 566 | hCoV-19/England/CAMC-A58AC5/2020\|EPI_ISL_596977\|20 | ···················· | ···················· | ···················· | ···················· | ···················· | ···················· |
| N | 567 | hCoV-19/England/CAMC-A58E32/2020\|EPI_ISL_596978\|20 | ···················· | ···················· | ···················· | ···················· | ···················· | ···················· |
| N | 568 | hCoV-19/England/CAMC-A583CA/2020\|EPI_ISL_596979\|20 | ···················· | ···················· | ···················· | ···················· | ···················· | ···················· |
| N | 569 | hCoV-19/England/CAMC-A5852E/2020\|EPI_ISL_596980\|20 | ···················· | ···················· | ···················· | ···················· | ···················· | ···················· |
| N | 570 | hCoV-19/England/CAMC-A58E14/2020\|EPI_ISL_596981\|20 | ···················· | ···················· | ···················· | ···················· | ···················· | ···················· |
| N | 571 | hCoV-19/Scotland/QEUH-A22578/2020\|EPI_ISL_599100\|2 | ···················· | ···················· | ···················· | ···················· | ···················· | ···················· |
| N | 572 | hCoV-19/Scotland/QEUH-A1FE37/2020\|EPI_ISL_599103\|2 | ···················· | ···················· | ···················· | ···················· | ···················· | ···················· |
| N | 573 | hCoV-19/Scotland/QEUH-A1F848/2020\|EPI_ISL_599104\|2 | ···················· | ···················· | ···················· | ···················· | ···················· | ···················· |
| N | 574 | hCoV-19/Scotland/QEUH-A1FBF4/2020\|EPI_ISL_599105\|2 | ···················· | ···················· | ···················· | ···················· | ···················· | ···················· |
| N | 575 | hCoV-19/Scotland/QEUH-A1F6F3/2020\|EPI_ISL_599107\|2 | ···················· | ···················· | ···················· | ···················· | ···················· | ···················· |
| N | 576 | hCoV-19/Scotland/QEUH-A1F8DF/2020\|EPI_ISL_599108\|2 | ···················· | ···················· | ···················· | ···················· | ···················· | ···················· |
| N | 577 | hCoV-19/Scotland/QEUH-A1FE0A/2020\|EPI_ISL_599109\|2 | ···················· | ···················· | ···················· | ···················· | ···················· | ···················· |
| N | 578 | hCoV-19/Scotland/QEUH-A1F893/2020\|EPI_ISL_599110\|2 | ···················· | ···················· | ···················· | ···················· | ···················· | ···················· |
| N | 579 | hCoV-19/Scotland/QEUH-A1F981/2020\|EPI_ISL_599111\|2 | ···················· | ···················· | ···················· | ···················· | ···················· | ···················· |
| N | 580 | hCoV-19/Scotland/QEUH-A1F602/2020\|EPI_ISL_599112\|2 | ···················· | ···················· | ···················· | ···················· | ···················· | ···················· |
| N | 581 | hCoV-19/Scotland/QEUH-A208BD/2020\|EPI_ISL_599113\|2 | ···················· | ···················· | ···················· | ···················· | ···················· | ···················· |
| N | 582 | hCoV-19/Scotland/QEUH-A1FD0D/2020\|EPI_ISL_599115\|2 | ···················· | ···················· | ···················· | ···················· | ···················· | ···················· |
| N | 583 | hCoV-19/Scotland/QEUH-A1FF8F/2020\|EPI_ISL_599116\|2 | ···················· | ···················· | ···················· | ···················· | ···················· | ···················· |
| N | 584 | hCoV-19/Scotland/QEUH-A1F918/2020\|EPI_ISL_599117\|2 | ···················· | ···················· | ···················· | ···················· | ···················· | ···················· |
| N | 585 | hCoV-19/Scotland/QEUH-A20228/2020\|EPI_ISL_599118\|2 | ···················· | ···················· | ···················· | ···················· | ···················· | ···················· |
| N | 586 | hCoV-19/Scotland/QEUH-A1FFAD/2020\|EPI_ISL_599120\|2 | ···················· | ···················· | ···················· | ···················· | ···················· | ···················· |
| N | 587 | hCoV-19/Scotland/QEUH-A1FB03/2020\|EPI_ISL_599121\|2 | ···················· | ···················· | ···················· | ···················· | ···················· | ···················· |
| N | 588 | hCoV-19/Scotland/QEUH-A1F990/2020\|EPI_ISL_599122\|2 | ···················· | ···················· | ···················· | ···················· | ···················· | ···················· |
| N | 589 | hCoV-19/Scotland/QEUH-A1F550/2020\|EPI_ISL_599123\|2 | ···················· | ···················· | ···················· | ···················· | ···················· | ···················· |
| N | 590 | hCoV-19/Scotland/QEUH-A1F5E7/2020\|EPI_ISL_599124\|2 | ···················· | ···················· | ···················· | ···················· | ···················· | ···················· |
| N | 591 | hCoV-19/Scotland/QEUH-A1FD49/2020\|EPI_ISL_599125\|2 | ···················· | ···················· | ···················· | ···················· | ···················· | ···················· |
| N | 592 | hCoV-19/Scotland/QEUH-A1F5C9/2020\|EPI_ISL_599126\|2 | ···················· | ···················· | ···················· | ···················· | ···················· | ···················· |
| N | 593 | hCoV-19/Scotland/QEUH-A22587/2020\|EPI_ISL_599127\|2 | ···················· | ···················· | ···················· | ···················· | ···················· | ···················· |
| N | 594 | hCoV-19/Scotland/QEUH-A1FA7F/2020\|EPI_ISL_599128\|2 | ···················· | ···················· | ···················· | ···················· | ···················· | ···················· |
| N | 595 | hCoV-19/Scotland/QEUH-A1F927/2020\|EPI_ISL_599129\|2 | ···················· | ···················· | ···················· | ···················· | ···················· | ···················· |
| N | 596 | hCoV-19/Scotland/QEUH-A1FABB/2020\|EPI_ISL_599130\|2 | ···················· | ···················· | ···················· | ···················· | ···················· | ···················· |
| N | 597 | hCoV-19/Scotland/QEUH-A20D09/2020\|EPI_ISL_599131\|2 | ···················· | ···················· | ···················· | ···················· | ···················· | ···················· |
| N | 598 | hCoV-19/Scotland/QEUH-A1F7C3/2020\|EPI_ISL_599133\|2 | ···················· | ···················· | ···················· | ···················· | ···················· | ···················· |
| N | 599 | hCoV-19/Scotland/QEUH-A20DCD/2020\|EPI_ISL_599134\|2 | ···················· | ···················· | ···················· | ···················· | ···················· | ···················· |
| N | 600 | hCoV-19/Scotland/QEUH-A1FEA0/2020\|EPI_ISL_599135\|2 | ···················· | ···················· | ···················· | ···················· | ···················· | ···················· |
| N | 601 | hCoV-19/Scotland/QEUH-A1FE19/2020\|EPI_ISL_599136\|2 | ···················· | ···················· | ···················· | ···················· | ···················· | ···················· |
| N | 602 | hCoV-19/Scotland/QEUH-A1FEBF/2020\|EPI_ISL_599137\|2 | ···················· | ···················· | ···················· | ···················· | ···················· | ···················· |
| N | 603 | hCoV-19/England/QEUH-A0B94C/2020\|EPI_ISL_599138\|20 | ···················· | ···················· | ···················· | ···················· | ····T··············· | ···················· |
| N | 604 | hCoV-19/Scotland/QEUH-A0B5B1/2020\|EPI_ISL_599160\|2 | ···················· | ···················· | ···················· | ···················· | ···················· | ···················· |
| N | 605 | hCoV-19/Scotland/QEUH-A0B89A/2020\|EPI_ISL_599161\|2 | ···················· | ···················· | ···················· | ···················· | ···················· | ···················· |
| N | 606 | hCoV-19/England/QEUH-A0BF4A/2020\|EPI_ISL_599162\|20 | ···················· | ···················· | ···················· | ···················· | ···················· | ···················· |
| N | 607 | hCoV-19/England/QEUH-A0BEA7/2020\|EPI_ISL_599163\|20 | ···················· | ···················· | ···················· | ···················· | ···················· | ···················· |
| N | 608 | hCoV-19/Wales/QEUH-A0BB46/2020\|EPI_ISL_599164\|2020 | ···················· | ···················· | ···················· | ···················· | ···················· | ···················· |
| N | 609 | hCoV-19/England/QEUH-A0B9D3/2020\|EPI_ISL_599167\|20 | ···················· | ···················· | ···················· | ···················· | ···················· | ···················· |
| N | 610 | hCoV-19/Scotland/QEUH-A0B496/2020\|EPI_ISL_599169\|2 | ···················· | ···················· | ···················· | ···················· | ···················· | ···················· |
| N | 611 | hCoV-19/England/QEUH-A0C820/2020\|EPI_ISL_599170\|20 | ···················· | ···················· | ···················· | ···················· | ···················· | ···················· |
| N | 612 | hCoV-19/Scotland/QEUH-A0B7CA/2020\|EPI_ISL_599171\|2 | ···················· | ···················· | ···················· | ···················· | ···················· | ···················· |
| N | 613 | hCoV-19/England/QEUH-A0AAFF/2020\|EPI_ISL_599172\|20 | ···················· | ···················· | ···················· | ···················· | ···················· | ···················· |
| N | 614 | hCoV-19/Scotland/QEUH-A0BC9E/2020\|EPI_ISL_599173\|2 | ···················· | ···················· | ···················· | ···················· | ···················· | ···················· |
| N | 615 | hCoV-19/England/QEUH-A0B0DE/2020\|EPI_ISL_599174\|20 | ···················· | ···················· | ···················· | ···················· | ···················· | ···················· |
| N | 616 | hCoV-19/England/QEUH-A0ACF9/2020\|EPI_ISL_599175\|20 | ···················· | ·A·················· | ···················· | ···················· | ···················· | ···················· |
| N | 617 | hCoV-19/Scotland/QEUH-A0AE4E/2020\|EPI_ISL_599176\|2 | ···················· | ···················· | ···················· | ···················· | ···················· | ···················· |
| N | 618 | hCoV-19/Scotland/QEUH-A0BC70/2020\|EPI_ISL_599177\|2 | ···················· | ···················· | ···················· | ···················· | ···················· | ···················· |
| N | 619 | hCoV-19/England/QEUH-A0B9E2/2020\|EPI_ISL_599178\|20 | ···················· | ···················· | ···················· | ···················· | ···················· | ···················· |
| N | 620 | hCoV-19/Scotland/QEUH-A0B5C0/2020\|EPI_ISL_599179\|2 | ···················· | ···················· | ···················· | ···················· | ···················· | ···················· |
| N | 621 | hCoV-19/Wales/QEUH-A0AD8D/2020\|EPI_ISL_599180\|2020 | ···················· | ···················· | ···················· | ···················· | ···················· | ···················· |
| N | 622 | hCoV-19/England/QEUH-A0B6FA/2020\|EPI_ISL_599181\|20 | ···················· | ···················· | ···················· | ···················· | ···················· | ···················· |
| N | 623 | hCoV-19/England/QEUH-A0B214/2020\|EPI_ISL_599182\|20 | ···················· | ···················· | ···················· | ···················· | ···················· | ···················· |
| N | 624 | hCoV-19/England/QEUH-A0B092/2020\|EPI_ISL_599184\|20 | ···················· | ···················· | ···················· | ···················· | ···················· | ···················· |
| N | 625 | hCoV-19/England/QEUH-A0BDE6/2020\|EPI_ISL_599185\|20 | ···················· | ···················· | ···················· | ···················· | ···················· | ···················· |
| N | 626 | hCoV-19/England/QEUH-A0BAD0/2020\|EPI_ISL_599186\|20 | ···················· | ···················· | ···················· | ···················· | ···················· | ···················· |
| N | 627 | hCoV-19/England/QEUH-A0AD14/2020\|EPI_ISL_599187\|20 | ···················· | ···················· | ···················· | ···················· | ······T············· | ···················· |
| N | 628 | hCoV-19/England/QEUH-A0BA58/2020\|EPI_ISL_599188\|20 | ···················· | ···················· | ···················· | ···················· | ···················· | ···················· |
| N | 629 | hCoV-19/Scotland/QEUH-A0BCF8/2020\|EPI_ISL_599189\|2 | ···················· | ···················· | ···················· | ···················· | ···················· | ···················· |
| N | 630 | hCoV-19/England/QEUH-A0AC17/2020\|EPI_ISL_599190\|20 | ···················· | ···················· | ···················· | ···················· | ···················· | ···················· |
| N | 631 | hCoV-19/Scotland/QEUH-A0B8F4/2020\|EPI_ISL_599191\|2 | ···················· | ···················· | ···················· | ···················· | ···················· | ···················· |
| N | 632 | hCoV-19/England/QEUH-A0B1EA/2020\|EPI_ISL_599192\|20 | ···················· | ···················· | ···················· | ···················· | ···················· | ···················· |

| | | | | | | | | |
|---|---|---|---|---|---|---|---|---|
| N | 633 | hCoV-19/England/QEUH-A0AC26/2020\|EPI_ISL_599193\|20 | ···················· | ···················· | ···················· | ···················· | ···················· | ···················· |
| N | 634 | hCoV-19/England/QEUH-A0BBBF/2020\|EPI_ISL_599195\|20 | ···················· | ···················· | ···················· | ···················· | ···················· | ···················· |
| N | 635 | hCoV-19/England/QEUH-A0BFFF/2020\|EPI_ISL_599196\|20 | ···················· | ···················· | ···················· | ···················· | ···················· | ···················· |
| N | 636 | hCoV-19/England/QEUH-A0C8F3/2020\|EPI_ISL_599197\|20 | ···················· | ·A·················· | ···················· | ···················· | ···················· | ···················· |
| N | 637 | hCoV-19/England/QEUH-A0B95B/2020\|EPI_ISL_599198\|20 | ···················· | ···················· | ···················· | ···················· | ···················· | ···················· |
| N | 638 | hCoV-19/England/QEUH-A0BFD1/2020\|EPI_ISL_599199\|20 | ···················· | ···················· | ···················· | ···················· | ···················· | ···················· |
| N | 639 | hCoV-19/USA/CA-QDX-1915/2020\|EPI_ISL_604200\|2020-0 | ···················· | ···················· | ···················· | ···················· | ···················· | ···················· |
| N | 640 | hCoV-19/USA/CA-QDX-1916/2020\|EPI_ISL_604201\|2020-0 | ···················· | ···················· | ···················· | ···················· | ···················· | ···················· |
| N | 641 | hCoV-19/USA/CA-QDX-1917/2020\|EPI_ISL_604202\|2020-0 | ···················· | ···················· | ···················· | ···················· | ···················· | ···················· |
| N | 642 | hCoV-19/USA/CA-QDX-1919/2020\|EPI_ISL_604203\|2020-0 | ···················· | ···················· | ···················· | ···················· | ···················· | ···················· |
| N | 643 | hCoV-19/USA/CA-QDX-1920/2020\|EPI_ISL_604204\|2020-0 | ···················· | ···················· | ···················· | ···················· | ···················· | ···················· |
| N | 644 | hCoV-19/USA/CA-QDX-1925/2020\|EPI_ISL_604205\|2020-0 | ···················· | ···················· | ···················· | ···················· | ···················· | ···················· |
| N | 645 | hCoV-19/USA/CA-QDX-1926/2020\|EPI_ISL_604206\|2020-0 | ···················· | ···················· | ···················· | ···················· | ···················· | ···················· |
| N | 646 | hCoV-19/USA/CA-QDX-1927/2020\|EPI_ISL_604207\|2020-0 | ···················· | ···················· | ···················· | ···················· | ···················· | ···················· |
| N | 647 | hCoV-19/USA/CA-QDX-1931/2020\|EPI_ISL_604208\|2020-0 | ···················· | ···················· | ···················· | ···················· | ···················· | ···················· |
| N | 648 | hCoV-19/USA/CA-QDX-1937/2020\|EPI_ISL_604209\|2020-0 | ···················· | ···················· | ···················· | ···················· | ···················· | ···················· |
| N | 649 | hCoV-19/USA/CA-QDX-1953/2020\|EPI_ISL_604211\|2020-0 | ···················· | ···················· | ···················· | ···················· | ···················· | ···················· |
| N | 650 | hCoV-19/USA/CA-QDX-1955/2020\|EPI_ISL_604212\|2020-0 | ···················· | ···················· | ···················· | ···················· | ···················· | ···················· |
| N | 651 | hCoV-19/USA/CA-QDX-1958/2020\|EPI_ISL_604213\|2020-0 | ···················· | ···················· | ···················· | ···················· | ···················· | ···················· |
| N | 652 | hCoV-19/USA/CA-QDX-1961/2020\|EPI_ISL_604214\|2020-0 | ···················· | ···················· | ···················· | ···················· | ···················· | ···················· |
| N | 653 | hCoV-19/USA/CA-QDX-1962/2020\|EPI_ISL_604215\|2020-0 | ···················· | ···················· | ···················· | ···················· | ···················· | ···················· |
| N | 654 | hCoV-19/USA/CA-QDX-1967/2020\|EPI_ISL_604216\|2020-0 | ···················· | ···················· | ···················· | ···················· | ···················· | ···················· |
| N | 655 | hCoV-19/USA/CA-QDX-1975/2020\|EPI_ISL_604217\|2020-0 | ···················· | ···················· | ···················· | ···················· | ···················· | ···················· |
| N | 656 | hCoV-19/USA/CA-QDX-1976/2020\|EPI_ISL_604218\|2020-0 | ···················· | ···················· | ···················· | ···················· | ···················· | ···················· |
| N | 657 | hCoV-19/USA/CA-QDX-1979/2020\|EPI_ISL_604219\|2020-0 | ···················· | ···················· | ···················· | ···················· | ···················· | ···················· |
| N | 658 | hCoV-19/USA/NY-QDX-1885/2020\|EPI_ISL_604240\|2020-0 | ···················· | ···················· | ···················· | ···················· | ···················· | ···················· |
| N | 659 | hCoV-19/USA/NY-QDX-1889/2020\|EPI_ISL_604241\|2020-0 | ···················· | ···················· | ···················· | ···················· | ···················· | ···················· |
| N | 660 | hCoV-19/USA/NY-QDX-1890/2020\|EPI_ISL_604242\|2020-0 | ···················· | ···················· | ···················· | ···················· | ···················· | ···················· |
| N | 661 | hCoV-19/USA/NY-QDX-1892/2020\|EPI_ISL_604243\|2020-0 | ···················· | ···················· | ···················· | ···················· | ···················· | ···················· |
| N | 662 | hCoV-19/USA/NY-QDX-1893/2020\|EPI_ISL_604244\|2020-0 | ···················· | ···················· | ···················· | ···················· | ···················· | ···················· |
| N | 663 | hCoV-19/USA/NY-QDX-1894/2020\|EPI_ISL_604245\|2020-0 | ···················· | ···················· | ···················· | ···················· | ···················· | ···················· |
| N | 664 | hCoV-19/USA/NY-QDX-1898/2020\|EPI_ISL_604246\|2020-0 | ···················· | ···················· | ···················· | ···················· | ···················· | ···················· |
| N | 665 | hCoV-19/USA/NY-QDX-1904/2020\|EPI_ISL_604247\|2020-0 | ···················· | ···················· | ···················· | ···················· | ···················· | ···················· |
| N | 666 | hCoV-19/USA/NY-QDX-1911/2020\|EPI_ISL_604249\|2020-0 | ···················· | ···················· | ···················· | ···················· | ···················· | ···················· |
| N | 667 | hCoV-19/USA/NY-QDX-1912/2020\|EPI_ISL_604250\|2020-0 | ···················· | ···················· | ···················· | ···················· | ···················· | ···················· |
| N | 668 | hCoV-19/USA/NY-QDX-1913/2020\|EPI_ISL_604251\|2020-0 | ···················· | ···················· | ···················· | ···················· | ···················· | ···················· |
| N | 669 | hCoV-19/USA/NY-QDX-1921/2020\|EPI_ISL_604252\|2020-0 | ···················· | ···················· | ···················· | ···················· | ···················· | ···················· |
| N | 670 | hCoV-19/USA/NY-QDX-1922/2020\|EPI_ISL_604253\|2020-0 | ···················· | ···················· | ···················· | ···················· | ···················· | ···················· |
| N | 671 | hCoV-19/USA/NY-QDX-1923/2020\|EPI_ISL_604254\|2020-0 | ···················· | ···················· | ···················· | ···················· | ···················· | ···················· |
| N | 672 | hCoV-19/USA/NY-QDX-1924/2020\|EPI_ISL_604255\|2020-0 | ···················· | ···················· | ···················· | ···················· | ···················· | ···················· |
| N | 673 | hCoV-19/USA/NY-QDX-1928/2020\|EPI_ISL_604256\|2020-0 | ···················· | ···················· | ···················· | ···················· | ···················· | ···················· |
| N | 674 | hCoV-19/USA/NY-QDX-1929/2020\|EPI_ISL_604257\|2020-0 | ···················· | ···················· | ···················· | ···················· | ···················· | ···················· |
| N | 675 | hCoV-19/USA/NY-QDX-1933/2020\|EPI_ISL_604258\|2020-0 | ···················· | ···················· | ···················· | ···················· | ···················· | ···················· |
| N | 676 | hCoV-19/USA/NY-QDX-1934/2020\|EPI_ISL_604259\|2020-0 | ···················· | ···················· | ···················· | ···················· | ···················· | ···················· |
| N | 677 | hCoV-19/USA/NY-QDX-1935/2020\|EPI_ISL_604260\|2020-0 | ···················· | ···················· | ···················· | ···················· | ···················· | ···················· |
| N | 678 | hCoV-19/USA/NY-QDX-1939/2020\|EPI_ISL_604261\|2020-0 | ···················· | ···················· | ···················· | ···················· | ···················· | ···················· |
| N | 679 | hCoV-19/USA/NY-QDX-1940/2020\|EPI_ISL_604262\|2020-0 | ···················· | ···················· | ···················· | ···················· | ···················· | ···················· |
| N | 680 | hCoV-19/USA/NY-QDX-1941/2020\|EPI_ISL_604263\|2020-0 | ···················· | ···················· | ···················· | ···················· | ···················· | ···················· |
| N | 681 | hCoV-19/USA/NY-QDX-1942/2020\|EPI_ISL_604264\|2020-0 | ···················· | ···················· | ···················· | ···················· | ···················· | ···················· |
| N | 682 | hCoV-19/USA/NY-QDX-1945/2020\|EPI_ISL_604265\|2020-0 | ···················· | ···················· | ···················· | ···················· | ···················· | ···················· |
| N | 683 | hCoV-19/USA/NY-QDX-1946/2020\|EPI_ISL_604266\|2020-0 | ···················· | ···················· | ···················· | ···················· | ···················· | ···················· |
| N | 684 | hCoV-19/USA/NY-QDX-1947/2020\|EPI_ISL_604267\|2020-0 | ···················· | ···················· | ···················· | ···················· | ···················· | ···················· |
| N | 685 | hCoV-19/USA/NY-QDX-1948/2020\|EPI_ISL_604268\|2020-0 | ···················· | ···················· | ···················· | ···················· | ···················· | ···················· |
| N | 686 | hCoV-19/USA/NY-QDX-1949/2020\|EPI_ISL_604269\|2020-0 | ···················· | ···················· | ···················· | ···················· | ···················· | ···················· |
| N | 687 | hCoV-19/USA/NY-QDX-1952/2020\|EPI_ISL_604270\|2020-0 | ···················· | ···················· | ···················· | ···················· | ···················· | ···················· |
| N | 688 | hCoV-19/USA/NY-QDX-1956/2020\|EPI_ISL_604271\|2020-0 | ···················· | ···················· | ···················· | ···················· | ···················· | ···················· |
| N | 689 | hCoV-19/USA/NY-QDX-1963/2020\|EPI_ISL_604272\|2020-0 | ···················· | ···················· | ···················· | ···················· | ···················· | ···················· |
| N | 690 | hCoV-19/USA/NY-QDX-1965/2020\|EPI_ISL_604273\|2020-0 | ···················· | ···················· | ···················· | ···················· | ···················· | ···················· |
| N | 691 | hCoV-19/USA/NY-QDX-1969/2020\|EPI_ISL_604274\|2020-0 | ···················· | ···················· | ···················· | ···················· | ···················· | ···················· |
| N | 692 | hCoV-19/USA/NY-QDX-1970/2020\|EPI_ISL_604275\|2020-0 | ···················· | ···················· | ···················· | ···················· | ···················· | ···················· |
| N | 693 | hCoV-19/USA/NY-QDX-1972/2020\|EPI_ISL_604276\|2020-0 | ···················· | ···················· | ···················· | ···················· | ···················· | ···················· |
| N | 694 | hCoV-19/USA/NY-QDX-1977/2020\|EPI_ISL_604277\|2020-0 | ···················· | ···················· | ···················· | ···················· | ···················· | ···················· |
| N | 695 | hCoV-19/USA/NY-QDX-1978/2020\|EPI_ISL_604278\|2020-0 | ···················· | ···················· | ···················· | ···················· | ···················· | ···················· |
| N | 696 | hCoV-19/USA/PA-QDX-1888/2020\|EPI_ISL_604279\|2020-0 | ···················· | ···················· | ···················· | ···················· | ···················· | ···················· |
| N | 697 | hCoV-19/USA/WI-UW-1626/2020\|EPI_ISL_605500\|2020-10 | ···················· | ···················· | ···················· | ···················· | ···················· | ···················· |
| N | 698 | hCoV-19/USA/WI-UW-1629/2020\|EPI_ISL_605503\|2020-09 | ···················· | ·A·················· | ···················· | ···················· | ···················· | ···················· |
| N | 699 | hCoV-19/USA/WI-UW-1634/2020\|EPI_ISL_605508\|2020-09 | ···················· | ···················· | ···················· | ···················· | ···················· | ···················· |
| N | 700 | hCoV-19/USA/WI-UW-1635/2020\|EPI_ISL_605509\|2020-09 | ···················· | ···················· | ···················· | ···················· | ···················· | ···················· |
| N | 701 | hCoV-19/USA/WI-UW-1636/2020\|EPI_ISL_605510\|2020-09 | ···················· | ···················· | ···················· | ···················· | ···················· | ···················· |
| N | 703 | hCoV-19/USA/WI-UW-1638/2020\|EPI_ISL_605512\|2020-09 | ···················· | ···················· | ···················· | ···················· | ···················· | ···················· |
| N | 704 | hCoV-19/USA/WI-UW-1639/2020\|EPI_ISL_605513\|2020-09 | ···················· | ···················· | ···················· | ···················· | ···················· | ···················· |
| N | 705 | hCoV-19/USA/WI-UW-1640/2020\|EPI_ISL_605514\|2020-09 | ···················· | ···················· | ···················· | ···················· | ···················· | ···················· |
| N | 707 | hCoV-19/USA/WI-UW-1642/2020\|EPI_ISL_605516\|2020-09 | ···················· | ···················· | ···················· | ···················· | ···················· | ···················· |
| N | 710 | hCoV-19/USA/WI-UW-1645/2020\|EPI_ISL_605519\|2020-09 | ···················· | ···················· | ···················· | ···················· | ···················· | ···················· |
| N | 712 | hCoV-19/USA/WI-UW-1647/2020\|EPI_ISL_605521\|2020-09 | ···················· | ···················· | ···················· | ···················· | ···················· | ···················· |
| N | 713 | hCoV-19/USA/WI-UW-1648/2020\|EPI_ISL_605522\|2020-09 | ···················· | ···················· | ···················· | ···················· | ···················· | ···················· |
| N | 714 | hCoV-19/USA/WI-UW-1649/2020\|EPI_ISL_605523\|2020-09 | ···················· | ···················· | ···················· | ···················· | ···················· | ···················· |
| N | 715 | hCoV-19/USA/WI-UW-1650/2020\|EPI_ISL_605524\|2020-09 | ···················· | ···················· | ···················· | ···················· | ···················· | ···················· |
| N | 716 | hCoV-19/USA/WI-UW-1651/2020\|EPI_ISL_605525\|2020-09 | ···················· | ···················· | ···················· | ···················· | ···················· | ···················· |
| N | 717 | hCoV-19/USA/WI-UW-1652/2020\|EPI_ISL_605526\|2020-09 | ···················· | ···················· | ···················· | ···················· | ···················· | ···················· |

| | | | | | | | | |
|---|---|---|---|---|---|---|---|---|
| N | 718 | hCoV-19/USA/WI-UW-1653/2020\|EPI_ISL_605527\|2020-09 | ···················· | ···················· | ···················· | ···················· | ···················· | ···················· |
| N | 719 | hCoV-19/USA/WI-UW-1654/2020\|EPI_ISL_605528\|2020-09 | ···················· | ···················· | ···················· | ···················· | ···················· | ···················· |
| N | 721 | hCoV-19/USA/WI-UW-1656/2020\|EPI_ISL_605530\|2020-09 | ···················· | ···················· | ···················· | ···················· | ···················· | ···················· |
| N | 722 | hCoV-19/USA/WI-UW-1657/2020\|EPI_ISL_605531\|2020-09 | ···················· | ···················· | ···················· | ···················· | ···················· | ···················· |
| N | 723 | hCoV-19/USA/WI-UW-1658/2020\|EPI_ISL_605532\|2020-09 | ···················· | ···················· | ···················· | ···················· | ···················· | ···················· |
| N | 724 | hCoV-19/USA/WI-UW-1659/2020\|EPI_ISL_605533\|2020-09 | ···················· | ···················· | ···················· | ···················· | ···················· | ···················· |
| N | 725 | hCoV-19/USA/WI-UW-1660/2020\|EPI_ISL_605534\|2020-09 | ···················· | ···················· | ···················· | ···················· | ···················· | ···················· |
| N | 728 | hCoV-19/USA/WI-UW-1663/2020\|EPI_ISL_605537\|2020-09 | ···················· | ···················· | ···················· | ···················· | ···················· | ···················· |
| N | 730 | hCoV-19/USA/WI-UW-1665/2020\|EPI_ISL_605539\|2020-09 | ···················· | ···················· | ···················· | ···················· | ···················· | ···················· |
| N | 731 | hCoV-19/USA/WI-UW-1666/2020\|EPI_ISL_605540\|2020-09 | ···················· | ···················· | ···················· | ···················· | ···················· | ···················· |
| N | 732 | hCoV-19/USA/WI-UW-1667/2020\|EPI_ISL_605541\|2020-09 | ···················· | ···················· | ···················· | ···················· | ···················· | ···················· |
| N | 734 | hCoV-19/USA/WI-UW-1669/2020\|EPI_ISL_605543\|2020-09 | ···················· | ···················· | ···················· | ···················· | ···················· | ···················· |
| N | 736 | hCoV-19/USA/WI-UW-1671/2020\|EPI_ISL_605545\|2020-09 | ···················· | ···················· | ···················· | ···················· | ···················· | ···················· |
| N | 737 | hCoV-19/USA/WI-UW-1672/2020\|EPI_ISL_605546\|2020-09 | ···················· | ···················· | ···················· | ···················· | ···················· | ···················· |
| N | 739 | hCoV-19/USA/WI-UW-1674/2020\|EPI_ISL_605548\|2020-09 | ···················· | ···················· | ···················· | ···················· | ···················· | ···················· |
| N | 740 | hCoV-19/USA/WI-UW-1675/2020\|EPI_ISL_605549\|2020-09 | ···················· | ···················· | ···················· | ···················· | ···················· | ···················· |
| N | 743 | hCoV-19/USA/WI-UW-1699/2020\|EPI_ISL_605572\|2020-09 | ···················· | ···················· | ···················· | ···················· | ···················· | ···················· |
| N | 746 | hCoV-19/USA/WI-UW-1702/2020\|EPI_ISL_605575\|2020-09 | ···················· | ···················· | ···················· | ···················· | ···················· | ···················· |
| N | 748 | hCoV-19/USA/WI-UW-1704/2020\|EPI_ISL_605577\|2020-09 | ···················· | ···················· | ···················· | ···················· | ···················· | ···················· |
| N | 749 | hCoV-19/USA/WI-UW-1705/2020\|EPI_ISL_605578\|2020-09 | ···················· | ···················· | ···················· | ···················· | ···················· | ···················· |
| N | 751 | hCoV-19/USA/WI-UW-1707/2020\|EPI_ISL_605580\|2020-09 | ···················· | ···················· | ···················· | ···················· | ···················· | ···················· |
| N | 753 | hCoV-19/USA/WI-UW-1709/2020\|EPI_ISL_605582\|2020-09 | ···················· | ···················· | ···················· | ···················· | ···················· | ···················· |
| N | 754 | hCoV-19/USA/WI-UW-1710/2020\|EPI_ISL_605583\|2020-09 | ···················· | ···················· | ···················· | ···················· | ···················· | ···················· |
| N | 757 | hCoV-19/USA/WI-UW-1714/2020\|EPI_ISL_605587\|2020-09 | ···················· | ···················· | ···················· | ···················· | ···················· | ···················· |
| N | 758 | hCoV-19/USA/WI-UW-1715/2020\|EPI_ISL_605588\|2020-09 | ···················· | ···················· | ···················· | ···················· | ···················· | ···················· |
| N | 767 | hCoV-19/USA/WI-UW-1725/2020\|EPI_ISL_605598\|2020-09 | ···················· | ···················· | ···················· | ···················· | ···················· | ···················· |
| N | 769 | hCoV-19/England/CAMC-A58D80/2020\|EPI_ISL_596983\|20 | ···················· | ···················· | ···················· | ···················· | ···················· | ···················· |
| N | 770 | hCoV-19/England/CAMC-A587BF/2020\|EPI_ISL_596985\|20 | ···················· | ···················· | ···················· | ···················· | ···················· | ···················· |
| N | 771 | hCoV-19/England/CAMC-A598F7/2020\|EPI_ISL_596986\|20 | ···················· | ···················· | ···················· | ···················· | ···················· | ···················· |
| N | 773 | hCoV-19/England/CAMC-A57FF4/2020\|EPI_ISL_596988\|20 | ···················· | ···················· | ···················· | ···················· | ···················· | ···················· |
| N | 774 | hCoV-19/England/CAMC-A58C65/2020\|EPI_ISL_596989\|20 | ···················· | ···················· | ···················· | ···················· | ···················· | ···················· |
| N | 775 | hCoV-19/England/CAMC-A583BB/2020\|EPI_ISL_596990\|20 | ···················· | ···················· | ···················· | ···················· | ···················· | ···················· |
| N | 776 | hCoV-19/England/CAMC-A58F6C/2020\|EPI_ISL_596991\|20 | ···················· | ···················· | ···················· | ···················· | ···················· | ···················· |
| N | 777 | hCoV-19/England/CAMC-A59332/2020\|EPI_ISL_596992\|20 | ···················· | ···················· | ···················· | ···················· | ···················· | ···················· |
| N | 778 | hCoV-19/England/CAMC-A583F7/2020\|EPI_ISL_596993\|20 | ···················· | ···················· | ···················· | ···················· | ···················· | ···················· |
| N | 779 | hCoV-19/England/CAMC-A585D3/2020\|EPI_ISL_596994\|20 | ···················· | ···················· | ···················· | ···················· | ···················· | ···················· |
| N | 780 | hCoV-19/England/CAMC-A58C92/2020\|EPI_ISL_596995\|20 | ···················· | ···················· | ···················· | ···················· | ···················· | ···················· |
| N | 781 | hCoV-19/England/CAMC-A5960C/2020\|EPI_ISL_596997\|20 | ···················· | ···················· | ···················· | ···················· | ···················· | ···················· |
| N | 782 | hCoV-19/England/CAMC-A59420/2020\|EPI_ISL_596998\|20 | ···················· | ···················· | ···················· | ···················· | ···················· | ···················· |
| N | 783 | hCoV-19/England/CAMC-A588DA/2020\|EPI_ISL_596999\|20 | ···················· | ···················· | ···················· | ···················· | ···················· | ···················· |
| N | 784 | hCoV-19/England/QEUH-A0BAA3/2020\|EPI_ISL_599139\|20 | ···················· | ···················· | ···················· | ···················· | ···················· | ···················· |
| N | 785 | hCoV-19/England/QEUH-A0B830/2020\|EPI_ISL_599140\|20 | ···················· | ···················· | ···················· | ···················· | ···················· | ···················· |
| N | 786 | hCoV-19/England/QEUH-A0AC53/2020\|EPI_ISL_599141\|20 | ···················· | ···················· | ···················· | ···················· | ···················· | ···················· |
| N | 787 | hCoV-19/England/QEUH-A0B250/2020\|EPI_ISL_599142\|20 | ···················· | ···················· | ···················· | ···················· | ······T············· | ···················· |
| N | 788 | hCoV-19/England/QEUH-A0B241/2020\|EPI_ISL_599143\|20 | ···················· | ···················· | ···················· | ···················· | ···················· | ···················· |
| N | 789 | hCoV-19/England/QEUH-A0B2F6/2020\|EPI_ISL_599144\|20 | ···················· | ·A·················· | ···················· | ···················· | ···················· | ···A················ |
| N | 790 | hCoV-19/Scotland/QEUH-A0AE11/2020\|EPI_ISL_599145\|2 | ···················· | ···················· | ···················· | ···················· | ···················· | ···················· |
| N | 791 | hCoV-19/England/QEUH-A0C84E/2020\|EPI_ISL_599146\|20 | ···················· | ·A·················· | ···················· | ···················· | ···················· | ···················· |
| N | 792 | hCoV-19/England/QEUH-A0B056/2020\|EPI_ISL_599147\|20 | ···················· | ···················· | ···················· | ···················· | ···················· | ···················· |
| N | 793 | hCoV-19/England/QEUH-A0B8C7/2020\|EPI_ISL_599148\|20 | ···················· | ···················· | ···················· | ···················· | ···················· | ···················· |
| N | 794 | hCoV-19/England/QEUH-A0B5A2/2020\|EPI_ISL_599149\|20 | ···················· | ···················· | ···················· | ···················· | ···················· | ···················· |
| N | 795 | hCoV-19/England/QEUH-A0B083/2020\|EPI_ISL_599150\|20 | ···················· | ···················· | ···················· | ···················· | ···················· | ···················· |
| N | 796 | hCoV-19/England/QEUH-A0BC07/2020\|EPI_ISL_599151\|20 | ···················· | ···················· | ···················· | ···················· | ···················· | ···················· |
| N | 797 | hCoV-19/England/QEUH-A0B144/2020\|EPI_ISL_599152\|20 | ···················· | ···················· | ···················· | ···················· | ···················· | ···················· |
| N | 798 | hCoV-19/England/QEUH-A0BDC8/2020\|EPI_ISL_599153\|20 | ···················· | ···················· | ···················· | ···················· | ···················· | ···················· |
| N | 799 | hCoV-19/England/QEUH-A0ACDB/2020\|EPI_ISL_599154\|20 | ···················· | ···················· | ···················· | ···················· | ···················· | ···················· |
| N | 800 | hCoV-19/England/QEUH-A0AF0F/2020\|EPI_ISL_599155\|20 | ···················· | ···················· | ···················· | ···················· | ···················· | ···················· |
| N | 801 | hCoV-19/England/QEUH-A0AC44/2020\|EPI_ISL_599156\|20 | ···················· | ···················· | ···················· | ···················· | ···················· | ···················· |
| N | 802 | hCoV-19/England/QEUH-A0AF3C/2020\|EPI_ISL_599157\|20 | ···················· | ···················· | ···················· | ···················· | ······T············· | ···················· |
| N | 803 | hCoV-19/England/QEUH-A0BE89/2020\|EPI_ISL_599158\|20 | ···················· | ···················· | ···················· | ···················· | ···················· | ···················· |
| N | 804 | hCoV-19/England/QEUH-A0C389/2020\|EPI_ISL_599159\|20 | ···················· | ···················· | ···················· | ···················· | ···················· | ···················· |
| N | 805 | hCoV-19/England/QEUH-A0B36C/2020\|EPI_ISL_599200\|20 | ···················· | ···················· | ···················· | ···················· | ······T············· | ···················· |
| N | 806 | hCoV-19/England/QEUH-A0ACAE/2020\|EPI_ISL_599202\|20 | ···················· | ···················· | ···················· | ···················· | ···················· | ···················· |
| N | 807 | hCoV-19/England/QEUH-A0C32F/2020\|EPI_ISL_599203\|20 | ···················· | ···················· | ···················· | ···················· | ···················· | ···················· |
| N | 808 | hCoV-19/England/QEUH-A0B7E8/2020\|EPI_ISL_599204\|20 | ···················· | ···················· | ···················· | ···················· | ···················· | ···················· |
| N | 809 | hCoV-19/Scotland/QEUH-A0B4C3/2020\|EPI_ISL_599206\|2 | ···················· | ···················· | ···················· | ···················· | ···················· | ···················· |
| N | 810 | hCoV-19/England/QEUH-A0B584/2020\|EPI_ISL_599207\|20 | ···················· | ···················· | ···················· | ···················· | ···················· | ···················· |
| N | 811 | hCoV-19/England/QEUH-A0B45A/2020\|EPI_ISL_599208\|20 | ···················· | ···················· | ···················· | ···················· | ···················· | ···················· |
| N | 812 | hCoV-19/Scotland/QEUH-A0BCDA/2020\|EPI_ISL_599209\|2 | ···················· | ···················· | ···················· | ···················· | ···················· | ···················· |
| N | 813 | hCoV-19/England/QEUH-A0C811/2020\|EPI_ISL_599210\|20 | ···················· | ·A·················· | ···················· | ···················· | ···················· | ···················· |
| N | 814 | hCoV-19/Scotland/QEUH-A0C33E/2020\|EPI_ISL_599211\|2 | ···················· | ···················· | ···················· | ···················· | ···················· | ···················· |
| N | 815 | hCoV-19/Scotland/QEUH-A0BD40/2020\|EPI_ISL_599212\|2 | ···················· | ···················· | ···················· | ···················· | ···················· | ···················· |
| N | 816 | hCoV-19/England/QEUH-A0B742/2020\|EPI_ISL_599213\|20 | ···················· | ···················· | ···················· | ···················· | ···················· | ···················· |
| N | 817 | hCoV-19/Scotland/QEUH-A0B8D6/2020\|EPI_ISL_599214\|2 | ···················· | ···················· | ···················· | ···················· | ···················· | ···················· |
| N | 818 | hCoV-19/Scotland/QEUH-A0B7D9/2020\|EPI_ISL_599215\|2 | ···················· | ···················· | ···················· | ···················· | ···················· | ···················· |
| N | 819 | hCoV-19/Scotland/QEUH-9FAB0F/2020\|EPI_ISL_600600\|2 | ···················· | ···················· | ···················· | ···················· | ···················· | ···················· |
| N | 820 | hCoV-19/Scotland/QEUH-9FB4A9/2020\|EPI_ISL_600601\|2 | ···················· | ···················· | ···················· | ···················· | ···················· | ···················· |
| N | 821 | hCoV-19/Scotland/QEUH-9FB245/2020\|EPI_ISL_600602\|2 | ···················· | ···················· | ···················· | ···················· | ···················· | ···················· |
| N | 822 | hCoV-19/Scotland/QEUH-9FA3BC/2020\|EPI_ISL_600604\|2 | ···················· | ···················· | ···················· | ···················· | ···················· | ···················· |
| N | 823 | hCoV-19/Scotland/QEUH-9FB60D/2020\|EPI_ISL_600606\|2 | ···················· | ···················· | ···················· | ···················· | ···················· | ···················· |

| | | | | | | | |
|---|---|---|---|---|---|---|---|
| N | 824 hCoV-19/Wales/QEUH-9FB500/2020\|EPI_ISL_600607\|2020 | ···················· | ···················· | ····················· | ···················· | ···················· | ···················· |
| N | 825 hCoV-19/Scotland/QEUH-9FA941/2020\|EPI_ISL_600608\|2 | ···················· | ···················· | ····················· | ···················· | ···················· | ···················· |
| N | 826 hCoV-19/Scotland/QEUH-9FB254/2020\|EPI_ISL_600609\|2 | ···················· | ···················· | ····················· | ···················· | ···················· | ···················· |
| N | 827 hCoV-19/Scotland/QEUH-9FA932/2020\|EPI_ISL_600610\|2 | ···················· | ···················· | ····················· | ···················· | ···················· | ···················· |
| N | 828 hCoV-19/Scotland/QEUH-9FB5C4/2020\|EPI_ISL_600611\|2 | ···················· | ···················· | ····················· | ···················· | ···················· | ···················· |
| N | 829 hCoV-19/Scotland/QEUH-9FA950/2020\|EPI_ISL_600612\|2 | ···················· | ···················· | ····················· | ···················· | ···················· | ···················· |
| N | 830 hCoV-19/Scotland/QEUH-9FB1DF/2020\|EPI_ISL_600613\|2 | ···················· | ···················· | ····················· | ···················· | ···················· | ···················· |
| N | 831 hCoV-19/Scotland/QEUH-9FB48B/2020\|EPI_ISL_600614\|2 | ···················· | ···················· | ····················· | ···················· | ···················· | ···················· |
| N | 832 hCoV-19/Scotland/QEUH-9FA4AA/2020\|EPI_ISL_600615\|2 | ···················· | ···················· | ····················· | ···················· | ···················· | ···················· |
| N | 833 hCoV-19/Scotland/QEUH-9FAAE4/2020\|EPI_ISL_600616\|2 | ···················· | ···················· | ····················· | ···················· | ···················· | ···················· |
| N | 834 hCoV-19/Scotland/QEUH-9FA96F/2020\|EPI_ISL_600617\|2 | ···················· | ···················· | ····················· | ···················· | ···················· | ···················· |
| N | 835 hCoV-19/Scotland/QEUH-9FB3CA/2020\|EPI_ISL_600618\|2 | ···················· | ···················· | ····················· | ···················· | ···················· | ···················· |
| N | 836 hCoV-19/Scotland/QEUH-9F4A17/2020\|EPI_ISL_600619\|2 | ···················· | ···················· | ····················· | ···················· | ···················· | ···················· |
| N | 837 hCoV-19/Scotland/QEUH-9F4A26/2020\|EPI_ISL_600620\|2 | ···················· | ···················· | ····················· | ···················· | ···················· | ···················· |
| N | 838 hCoV-19/Scotland/QEUH-9F65E7/2020\|EPI_ISL_600621\|2 | ···················· | ···················· | ····················· | ···················· | ···················· | ···················· |
| N | 839 hCoV-19/Scotland/QEUH-9F5A8F/2020\|EPI_ISL_600622\|2 | ···················· | ···················· | ····················· | ···················· | ···················· | ···················· |
| N | 840 hCoV-19/Scotland/QEUH-9F6954/2020\|EPI_ISL_600623\|2 | ···················· | ···················· | ····················· | ···················· | ···················· | ···················· |
| N | 841 hCoV-19/Scotland/QEUH-9F472F/2020\|EPI_ISL_600624\|2 | ···················· | ···················· | ····················· | ···················· | ···················· | ···················· |
| N | 842 hCoV-19/Scotland/QEUH-9F66A8/2020\|EPI_ISL_600625\|2 | ···················· | ···················· | ····················· | ···················· | ···················· | ···················· |
| N | 843 hCoV-19/Scotland/QEUH-9F46C8/2020\|EPI_ISL_600626\|2 | ···················· | ···················· | ····················· | ···················· | ···················· | ···················· |
| N | 844 hCoV-19/Scotland/QEUH-9F671E/2020\|EPI_ISL_600627\|2 | ···················· | ···················· | ····················· | ···················· | ···················· | ···················· |
| N | 845 hCoV-19/Scotland/QEUH-9F6B4F/2020\|EPI_ISL_600628\|2 | ···················· | ···················· | ····················· | ···················· | ···················· | ···················· |
| N | 846 hCoV-19/Scotland/QEUH-9F6972/2020\|EPI_ISL_600629\|2 | ···················· | ···················· | ····················· | ···················· | ···················· | ···················· |
| N | 847 hCoV-19/Scotland/QEUH-9F6963/2020\|EPI_ISL_600630\|2 | ···················· | ···················· | ····················· | ···················· | ···················· | ···················· |
| N | 848 hCoV-19/Scotland/QEUH-9F5E29/2020\|EPI_ISL_600631\|2 | ···················· | ···················· | ····················· | ···················· | ···················· | ···················· |
| N | 849 hCoV-19/Scotland/QEUH-9F46D7/2020\|EPI_ISL_600632\|2 | ···················· | ···················· | ····················· | ···················· | ···················· | ···················· |
| N | 850 hCoV-19/Scotland/QEUH-9F6ACA/2020\|EPI_ISL_600633\|2 | ···················· | ···················· | ····················· | ···················· | ···················· | ···················· |
| N | 851 hCoV-19/Scotland/QEUH-9F6778/2020\|EPI_ISL_600634\|2 | ···················· | ···················· | ····················· | ···················· | ···················· | ···················· |
| N | 852 hCoV-19/Scotland/QEUH-9F4419/2020\|EPI_ISL_600636\|2 | ···················· | ···················· | ····················· | ···················· | ···················· | ···················· |
| N | 853 hCoV-19/Scotland/QEUH-9F4B7E/2020\|EPI_ISL_600637\|2 | ···················· | ···················· | ····················· | ···················· | ···················· | ···················· |
| N | 854 hCoV-19/Scotland/QEUH-9F4C8A/2020\|EPI_ISL_600638\|2 | ···················· | ···················· | ····················· | ···················· | ·····T·············· | ···················· |
| N | 855 hCoV-19/Scotland/QEUH-9F656F/2020\|EPI_ISL_600679\|2 | ···················· | ···················· | ····················· | ···················· | ···················· | ···················· |
| N | 856 hCoV-19/Scotland/QEUH-9F69EB/2020\|EPI_ISL_600680\|2 | ···················· | ···················· | ····················· | ···················· | ···················· | ···················· |
| N | 857 hCoV-19/Scotland/QEUH-9F4525/2020\|EPI_ISL_600681\|2 | ···················· | ···················· | ····················· | ···················· | ···················· | ···················· |
| N | 858 hCoV-19/Scotland/QEUH-9F4789/2020\|EPI_ISL_600682\|2 | ···················· | ···················· | ····················· | ···················· | ···················· | ···················· |
| N | 859 hCoV-19/Scotland/QEUH-9F6602/2020\|EPI_ISL_600683\|2 | ···················· | ···················· | ····················· | ···················· | ···················· | ···················· |
| N | 860 hCoV-19/Scotland/QEUH-9F5FBD/2020\|EPI_ISL_600684\|2 | ···················· | ···················· | ····················· | ···················· | ···················· | ···················· |
| N | 861 hCoV-19/Scotland/QEUH-9F6ABB/2020\|EPI_ISL_600685\|2 | ···················· | ···················· | ····················· | ···················· | ···················· | ···················· |
| N | 862 hCoV-19/Scotland/QEUH-9F6699/2020\|EPI_ISL_600686\|2 | ···················· | ···················· | ····················· | ···················· | ···················· | ···················· |
| N | 863 hCoV-19/Scotland/QEUH-9F4464/2020\|EPI_ISL_600687\|2 | ···················· | ···················· | ····················· | ···················· | ···················· | ···················· |
| N | 864 hCoV-19/Scotland/QEUH-9F69FA/2020\|EPI_ISL_600689\|2 | ···················· | ···················· | ····················· | ···················· | ···················· | ···················· |
| N | 865 hCoV-19/Scotland/QEUH-9F46F5/2020\|EPI_ISL_600690\|2 | ···················· | ···················· | ····················· | ···················· | ···················· | ···················· |
| N | 866 hCoV-19/Scotland/QEUH-9F4455/2020\|EPI_ISL_600691\|2 | ···················· | ···················· | ····················· | ···················· | ···················· | ···················· |
| N | 867 hCoV-19/Scotland/QEUH-9F4428/2020\|EPI_ISL_600692\|2 | ···················· | ···················· | ····················· | ···················· | ···················· | ···················· |
| N | 868 hCoV-19/Scotland/QEUH-9F5DE0/2020\|EPI_ISL_600693\|2 | ···················· | ···················· | ····················· | ···················· | ···················· | ···················· |
| N | 869 hCoV-19/Scotland/QEUH-9F6532/2020\|EPI_ISL_600694\|2 | ···················· | ···················· | ····················· | ···················· | ···················· | ···················· |
| N | 870 hCoV-19/Scotland/QEUH-9F4A71/2020\|EPI_ISL_600695\|2 | ···················· | ···················· | ····················· | ···················· | ···················· | ···················· |
| N | 871 hCoV-19/Scotland/QEUH-9F4C6C/2020\|EPI_ISL_600696\|2 | ···················· | ···················· | ····················· | ···················· | ···················· | ···················· |
| N | 872 hCoV-19/Scotland/QEUH-9F4AEA/2020\|EPI_ISL_600697\|2 | ···················· | ···················· | ····················· | ···················· | ···················· | ···················· |
| N | 873 hCoV-19/Scotland/QEUH-9F5AAD/2020\|EPI_ISL_600698\|2 | ···················· | ···················· | ····················· | ···················· | ···················· | ···················· |
| N | 874 hCoV-19/Scotland/QEUH-9F4A53/2020\|EPI_ISL_600699\|2 | ···················· | ···················· | ····················· | ···················· | ···················· | ···················· |
| N | 875 hCoV-19/Scotland/QEUH-9CEAE9/2020\|EPI_ISL_601900\|2 | ···················· | ···················· | ····················· | ···················· | ···················· | ···················· |
| N | 876 hCoV-19/Scotland/QEUH-9CE5E8/2020\|EPI_ISL_601901\|2 | ···················· | ···················· | ····················· | ···················· | ···················· | ···················· |
| N | 877 hCoV-19/Scotland/QEUH-9CECD4/2020\|EPI_ISL_601902\|2 | ···················· | ···················· | ····················· | ···················· | ···················· | ···················· |
| N | 878 hCoV-19/Scotland/QEUH-9CE287/2020\|EPI_ISL_601903\|2 | ···················· | ···················· | ····················· | ···················· | ···················· | ···················· |
| N | 879 hCoV-19/Scotland/QEUH-9CEE29/2020\|EPI_ISL_601904\|2 | ···················· | ···················· | ····················· | ···················· | ···················· | ···················· |
| N | 880 hCoV-19/Scotland/QEUH-9CF347/2020\|EPI_ISL_601905\|2 | ···················· | ···················· | ····················· | ···················· | ···················· | ···················· |
| N | 881 hCoV-19/Scotland/QEUH-9CE788/2020\|EPI_ISL_601906\|2 | ···················· | ···················· | ····················· | ···················· | ···················· | ···················· |
| N | 882 hCoV-19/Scotland/QEUH-9CE269/2020\|EPI_ISL_601907\|2 | ···················· | ···················· | ····················· | ···················· | ···················· | ···················· |
| N | 883 hCoV-19/Scotland/QEUH-9CEEDE/2020\|EPI_ISL_601908\|2 | ···················· | ···················· | ····················· | ···················· | ···················· | ···················· |
| N | 884 hCoV-19/Scotland/QEUH-9CE928/2020\|EPI_ISL_601909\|2 | ···················· | ···················· | ····················· | ···················· | ···················· | ···················· |
| N | 885 hCoV-19/Scotland/QEUH-9CF30B/2020\|EPI_ISL_601910\|2 | ···················· | ···················· | ····················· | ···················· | ···················· | ···················· |
| N | 886 hCoV-19/Scotland/QEUH-9CF7B4/2020\|EPI_ISL_601911\|2 | ···················· | ···················· | ····················· | ···················· | ···················· | ···················· |
| N | 887 hCoV-19/Scotland/QEUH-9CE621/2020\|EPI_ISL_601912\|2 | ···················· | ···················· | ····················· | ···················· | ···················· | ···················· |
| N | 888 hCoV-19/Scotland/QEUH-9CE9DD/2020\|EPI_ISL_601913\|2 | ···················· | ···················· | ····················· | ···················· | ···················· | ···················· |
| N | 889 hCoV-19/Scotland/QEUH-9CE612/2020\|EPI_ISL_601914\|2 | ···················· | ···················· | ····················· | ···················· | ···T················ | ···················· |
| N | 890 hCoV-19/Scotland/QEUH-9CEFCC/2020\|EPI_ISL_601915\|2 | ···················· | ·····C·············· | ····················· | ···················· | ···················· | ···················· |
| N | 891 hCoV-19/Scotland/QEUH-9CEF35/2020\|EPI_ISL_601916\|2 | ···················· | ···················· | ····················· | ···················· | ···················· | ···················· |
| N | 892 hCoV-19/Scotland/QEUH-9CEA52/2020\|EPI_ISL_601917\|2 | ···················· | ···················· | ····················· | ···················· | ···················· | ···················· |
| N | 893 hCoV-19/Scotland/QEUH-9CE409/2020\|EPI_ISL_601918\|2 | ···················· | ···················· | ····················· | ···················· | ···················· | ···················· |
| N | 894 hCoV-19/Scotland/QEUH-9CE90A/2020\|EPI_ISL_601919\|2 | ···················· | ···················· | ····················· | ···················· | ···················· | ···················· |
| N | 895 hCoV-19/Scotland/QEUH-9CEC5C/2020\|EPI_ISL_601920\|2 | ···················· | ···················· | ····················· | ···················· | ···················· | ···················· |
| N | 896 hCoV-19/Scotland/QEUH-9CEEB0/2020\|EPI_ISL_601921\|2 | ···················· | ···················· | ····················· | ···················· | ···················· | ···················· |
| N | 897 hCoV-19/Scotland/QEUH-9CF0D7/2020\|EPI_ISL_601922\|2 | ···················· | ···················· | ····················· | ···················· | ···················· | ···················· |
| N | 898 hCoV-19/Scotland/QEUH-9CF4BD/2020\|EPI_ISL_601923\|2 | ···················· | ···················· | ····················· | ···················· | ···················· | ···················· |
| N | 899 hCoV-19/Scotland/QEUH-9CF2FF/2020\|EPI_ISL_601925\|2 | ···················· | ···················· | ····················· | ···················· | ···················· | ···················· |
| N | 900 hCoV-19/Scotland/QEUH-9CE2A5/2020\|EPI_ISL_601926\|2 | ···················· | ···················· | ····················· | ···················· | ···················· | ···················· |
| N | 901 hCoV-19/Scotland/QEUH-9CF620/2020\|EPI_ISL_601927\|2 | ···················· | ···················· | ····················· | ···················· | ···················· | ···················· |
| N | 902 hCoV-19/Scotland/QEUH-9CEAF8/2020\|EPI_ISL_601930\|2 | ···················· | ···················· | ····················· | ···················· | ···················· | ···················· |
| N | 903 hCoV-19/Scotland/QEUH-9CF6E4/2020\|EPI_ISL_601931\|2 | ···················· | ···················· | ····················· | ···················· | ···················· | ···················· |

| | | | | | | | | |
|---|---|---|---|---|---|---|---|---|
| N | 904 | hCoV-19/Scotland/QEUH-9CEF62/2020\|EPI_ISL_601932\|2 | ···················· | ···················· | ····················· | ···················· | ···················· | ···················· |
| N | 905 | hCoV-19/Scotland/QEUH-9CE630/2020\|EPI_ISL_601933\|2 | ···················· | ···················· | ····················· | ···················· | ···················· | ···················· |
| N | 906 | hCoV-19/Scotland/QEUH-9CE6A9/2020\|EPI_ISL_601934\|2 | ···················· | ···················· | ····················· | ···················· | ···················· | ···················· |
| N | 907 | hCoV-19/Scotland/QEUH-9CE2D2/2020\|EPI_ISL_601935\|2 | ···················· | ···················· | ····················· | ···················· | ···················· | ···················· |
| N | 908 | hCoV-19/Scotland/QEUH-9CF6D5/2020\|EPI_ISL_601936\|2 | ···················· | ···················· | ····················· | ···················· | ···················· | ···················· |
| N | 909 | hCoV-19/Scotland/QEUH-9CE551/2020\|EPI_ISL_601937\|2 | ···················· | ···················· | ····················· | ···················· | ···················· | ···················· |
| N | 910 | hCoV-19/Scotland/QEUH-9CF4F9/2020\|EPI_ISL_601938\|2 | ···················· | ···················· | ····················· | ···················· | ···················· | ···················· |
| N | 911 | hCoV-19/Scotland/QEUH-9CEA16/2020\|EPI_ISL_601939\|2 | ···················· | ···················· | ····················· | ···················· | ···················· | ···················· |
| N | 912 | hCoV-19/Scotland/QEUH-9CE991/2020\|EPI_ISL_601940\|2 | ···················· | ···················· | ····················· | ···················· | ···················· | ···················· |
| N | 913 | hCoV-19/Scotland/QEUH-9CEEFC/2020\|EPI_ISL_601941\|2 | ···················· | ···················· | ····················· | ···················· | ···················· | ···················· |
| N | 914 | hCoV-19/Scotland/QEUH-9CF81B/2020\|EPI_ISL_601942\|2 | ···················· | ···················· | ····················· | ···················· | ···················· | ···················· |
| N | 915 | hCoV-19/Scotland/QEUH-9CF6C6/2020\|EPI_ISL_601943\|2 | ···················· | ···················· | ····················· | ···················· | ···················· | ···················· |
| N | 916 | hCoV-19/Scotland/QEUH-9CF2E0/2020\|EPI_ISL_601944\|2 | ···················· | ···················· | ····················· | ···················· | ···················· | ···················· |
| N | 917 | hCoV-19/Scotland/QEUH-9CE5D9/2020\|EPI_ISL_601945\|2 | ···················· | ···················· | ····················· | ···················· | ···················· | ···················· |
| N | 918 | hCoV-19/Scotland/QEUH-9CEF9F/2020\|EPI_ISL_601947\|2 | ···················· | ···················· | ····················· | ···················· | ···················· | ···················· |
| N | 919 | hCoV-19/Scotland/QEUH-9CE445/2020\|EPI_ISL_601948\|2 | ···················· | ···················· | ····················· | ···················· | ······T············· | ···················· |
| N | 920 | hCoV-19/Scotland/QEUH-9CE8FE/2020\|EPI_ISL_601949\|2 | ···················· | ···················· | ····················· | ···················· | ···················· | ···················· |
| N | 921 | hCoV-19/Scotland/QEUH-9CF7E1/2020\|EPI_ISL_601951\|2 | ···················· | ···················· | ····················· | ···················· | ···················· | ···················· |
| N | 922 | hCoV-19/Scotland/QEUH-9CEFF9/2020\|EPI_ISL_601952\|2 | ···················· | ···················· | ····················· | ···················· | ···················· | ···················· |
| N | 923 | hCoV-19/Scotland/QEUH-9CEEED/2020\|EPI_ISL_601953\|2 | ···················· | ···················· | ····················· | ···················· | ···················· | ···················· |
| N | 924 | hCoV-19/Scotland/QEUH-9CF383/2020\|EPI_ISL_601954\|2 | ···················· | ···················· | ····················· | ···················· | ···················· | ···················· |
| N | 925 | hCoV-19/Scotland/QEUH-9CE6B8/2020\|EPI_ISL_601955\|2 | ···················· | ···················· | ····················· | ···················· | ···················· | ···················· |
| N | 926 | hCoV-19/Scotland/QEUH-9CE955/2020\|EPI_ISL_601956\|2 | ···················· | ···················· | ····················· | ···················· | ···················· | ···················· |
| N | 927 | hCoV-19/Scotland/QEUH-9CEACB/2020\|EPI_ISL_601957\|2 | ···················· | ···················· | ····················· | ···················· | ···················· | ···················· |
| N | 928 | hCoV-19/Scotland/QEUH-9CF6A8/2020\|EPI_ISL_601958\|2 | ···················· | ···················· | ····················· | ···················· | ···················· | ···················· |
| N | 929 | hCoV-19/Scotland/QEUH-9CE2E1/2020\|EPI_ISL_601959\|2 | ···················· | ·A·······A·········· | ····················· | ···················· | ···················· | ···················· |
| N | 930 | hCoV-19/Scotland/QEUH-9CF68A/2020\|EPI_ISL_601960\|2 | ···················· | ···················· | ····················· | ···················· | ···················· | ···················· |
| N | 931 | hCoV-19/Scotland/QEUH-9CEE47/2020\|EPI_ISL_601961\|2 | ···················· | ···················· | ····················· | ···················· | ···················· | ···················· |
| N | 932 | hCoV-19/Scotland/QEUH-9CF374/2020\|EPI_ISL_601962\|2 | ···················· | ···················· | ····················· | ···················· | ···················· | ···················· |
| N | 933 | hCoV-19/Scotland/QEUH-9CF505/2020\|EPI_ISL_601963\|2 | ···················· | ···················· | ····················· | ···················· | ···················· | ···················· |
| N | 934 | hCoV-19/Scotland/QEUH-9CEE38/2020\|EPI_ISL_601964\|2 | ···················· | ···················· | ····················· | ···················· | ···················· | ···················· |
| N | 935 | hCoV-19/Scotland/QEUH-9CE3DF/2020\|EPI_ISL_601965\|2 | ···················· | ···················· | ····················· | ···················· | ···················· | ···················· |
| N | 936 | hCoV-19/Scotland/QEUH-9CF699/2020\|EPI_ISL_601966\|2 | ···················· | ···················· | ····················· | ···················· | ···················· | ···················· |
| N | 937 | hCoV-19/Scotland/QEUH-9CEA8F/2020\|EPI_ISL_601967\|2 | ···················· | ···················· | ····················· | ···················· | ···················· | ···················· |
| N | 938 | hCoV-19/Scotland/QEUH-9CEA70/2020\|EPI_ISL_601968\|2 | ···················· | ···················· | ····················· | ···················· | ···················· | ···················· |
| N | 939 | hCoV-19/Scotland/QEUH-9CEA9E/2020\|EPI_ISL_601969\|2 | ···················· | ···················· | ····················· | ···················· | ···················· | ···················· |
| N | 940 | hCoV-19/USA/UT-QDX-1844/2020\|EPI_ISL_604160\|2020-0 | ···················· | ···················· | ····················· | ···················· | ···················· | ···················· |
| N | 941 | hCoV-19/USA/UT-QDX-1845/2020\|EPI_ISL_604161\|2020-0 | ···················· | ···················· | ····················· | ···················· | ···················· | ···················· |
| N | 942 | hCoV-19/USA/UT-QDX-1846/2020\|EPI_ISL_604162\|2020-0 | ···················· | ···················· | ····················· | ···················· | ···················· | ···················· |
| N | 943 | hCoV-19/USA/UT-QDX-1847/2020\|EPI_ISL_604163\|2020-0 | ···················· | ···················· | ····················· | ···················· | ···················· | ···················· |
| N | 944 | hCoV-19/USA/UT-QDX-1848/2020\|EPI_ISL_604164\|2020-0 | ···················· | ···················· | ····················· | ···················· | ···················· | ···················· |
| N | 945 | hCoV-19/USA/UT-QDX-1849/2020\|EPI_ISL_604165\|2020-0 | ···················· | ···················· | ····················· | ···················· | ···················· | ···················· |
| N | 946 | hCoV-19/USA/UT-QDX-1850/2020\|EPI_ISL_604166\|2020-0 | ···················· | ···················· | ····················· | ···················· | ···················· | ···················· |
| N | 947 | hCoV-19/USA/UT-QDX-1851/2020\|EPI_ISL_604167\|2020-0 | ···················· | ···················· | ····················· | ···················· | ···················· | ···················· |
| N | 948 | hCoV-19/USA/UT-QDX-1852/2020\|EPI_ISL_604168\|2020-0 | ···················· | ···················· | ····················· | ···················· | ···················· | ···················· |
| N | 949 | hCoV-19/USA/UT-QDX-1853/2020\|EPI_ISL_604169\|2020-0 | ···················· | ···················· | ····················· | ···················· | ···················· | ···················· |
| N | 950 | hCoV-19/USA/UT-QDX-1854/2020\|EPI_ISL_604170\|2020-0 | ···················· | ···················· | ····················· | ···················· | ···················· | ···················· |
| N | 951 | hCoV-19/USA/UT-QDX-1856/2020\|EPI_ISL_604171\|2020-0 | ···················· | ···················· | ····················· | ···················· | ···················· | ···················· |
| N | 952 | hCoV-19/USA/UT-QDX-1858/2020\|EPI_ISL_604172\|2020-0 | ···················· | ···················· | ····················· | ···················· | ···················· | ···················· |
| N | 953 | hCoV-19/USA/UT-QDX-1859/2020\|EPI_ISL_604173\|2020-0 | ···················· | ···················· | ····················· | ···················· | ···················· | ···················· |
| N | 954 | hCoV-19/USA/UT-QDX-1860/2020\|EPI_ISL_604174\|2020-0 | ···················· | ···················· | ····················· | ···················· | ···················· | ···················· |
| N | 955 | hCoV-19/USA/UT-QDX-1861/2020\|EPI_ISL_604175\|2020-0 | ···················· | ···················· | ····················· | ···················· | ···················· | ···················· |
| N | 956 | hCoV-19/USA/UT-QDX-1863/2020\|EPI_ISL_604176\|2020-0 | ···················· | ···················· | ····················· | ···················· | ···················· | ···················· |
| N | 957 | hCoV-19/USA/UT-QDX-1864/2020\|EPI_ISL_604177\|2020-0 | ···················· | ···················· | ····················· | ···················· | ···················· | ···················· |
| N | 958 | hCoV-19/USA/UT-QDX-1865/2020\|EPI_ISL_604178\|2020-0 | ···················· | ···················· | ····················· | ···················· | ···················· | ···················· |
| N | 959 | hCoV-19/USA/UT-QDX-1866/2020\|EPI_ISL_604179\|2020-0 | ···················· | ···················· | ····················· | ···················· | ···················· | ···················· |
| N | 960 | hCoV-19/USA/UT-QDX-1867/2020\|EPI_ISL_604180\|2020-0 | ···················· | ···················· | ····················· | ···················· | ···················· | ···················· |
| N | 961 | hCoV-19/USA/UT-QDX-1868/2020\|EPI_ISL_604181\|2020-0 | ···················· | ···················· | ····················· | ···················· | ···················· | ···················· |
| N | 962 | hCoV-19/USA/UT-QDX-1870/2020\|EPI_ISL_604183\|2020-0 | ···················· | ···················· | ····················· | ···················· | ···················· | ···················· |
| N | 963 | hCoV-19/USA/AZ-QDX-1897/2020\|EPI_ISL_604184\|2020-0 | ···················· | ···················· | ····················· | ···················· | ···················· | ···················· |
| N | 964 | hCoV-19/USA/CA-QDX-1871/2020\|EPI_ISL_604185\|2020-0 | ···················· | ···················· | ····················· | ···················· | ···················· | ···················· |
| N | 965 | hCoV-19/USA/CA-QDX-1872/2020\|EPI_ISL_604186\|2020-0 | ···················· | ···················· | ····················· | ···················· | ···················· | ···················· |
| N | 966 | hCoV-19/USA/CA-QDX-1873/2020\|EPI_ISL_604187\|2020-0 | ···················· | ···················· | ····················· | ···················· | ···················· | ···················· |
| N | 967 | hCoV-19/USA/CA-QDX-1875/2020\|EPI_ISL_604188\|2020-0 | ···················· | ···················· | ····················· | ···················· | ···················· | ···················· |
| N | 968 | hCoV-19/USA/CA-QDX-1879/2020\|EPI_ISL_604189\|2020-0 | ···················· | ···················· | ····················· | ···················· | ···················· | ···················· |
| N | 969 | hCoV-19/USA/CA-QDX-1881/2020\|EPI_ISL_604190\|2020-0 | ···················· | ···················· | ····················· | ···················· | ···················· | ···················· |
| N | 970 | hCoV-19/USA/CA-QDX-1882/2020\|EPI_ISL_604191\|2020-0 | ···················· | ···················· | ····················· | ···················· | ···················· | ···················· |
| N | 971 | hCoV-19/USA/CA-QDX-1891/2020\|EPI_ISL_604192\|2020-0 | ···················· | ···················· | ····················· | ···················· | ···················· | ···················· |
| N | 972 | hCoV-19/USA/CA-QDX-1895/2020\|EPI_ISL_604193\|2020-0 | ···················· | ···················· | ····················· | ···················· | ···················· | ···················· |
| N | 973 | hCoV-19/USA/CA-QDX-1899/2020\|EPI_ISL_604194\|2020-0 | ···················· | ···················· | ····················· | ···················· | ···················· | ···················· |
| N | 974 | hCoV-19/USA/CA-QDX-1905/2020\|EPI_ISL_604196\|2020-0 | ···················· | ···················· | ····················· | ···················· | ···················· | ···················· |
| N | 975 | hCoV-19/USA/CA-QDX-1907/2020\|EPI_ISL_604197\|2020-0 | ···················· | ···················· | ····················· | ···················· | ···················· | ···················· |
| N | 976 | hCoV-19/USA/CA-QDX-1910/2020\|EPI_ISL_604198\|2020-0 | ···················· | ···················· | ····················· | ···················· | ···················· | ···················· |
| N | 977 | hCoV-19/USA/CA-QDX-1914/2020\|EPI_ISL_604199\|2020-0 | ···················· | ···················· | ····················· | ···················· | ···················· | ···················· |
| N | 978 | hCoV-19/USA/CO-QDX-1903/2020\|EPI_ISL_604220\|2020-0 | ···················· | ···················· | ····················· | ···················· | ···················· | ···················· |
| N | 979 | hCoV-19/USA/CO-QDX-1932/2020\|EPI_ISL_604221\|2020-0 | ···················· | ···················· | ····················· | ···················· | ···················· | ···················· |
| N | 980 | hCoV-19/USA/GA-QDX-1938/2020\|EPI_ISL_604222\|2020-0 | ···················· | ···················· | ····················· | ···················· | ···················· | ···················· |
| N | 981 | hCoV-19/USA/IN-QDX-1973/2020\|EPI_ISL_604223\|2020-0 | ···················· | ···················· | ····················· | ···················· | ···················· | ···················· |
| N | 982 | hCoV-19/USA/LA-QDX-1918/2020\|EPI_ISL_604224\|2020-0 | ···················· | ···················· | ····················· | ···················· | ···················· | ···················· |
| N | 983 | hCoV-19/USA/MI-QDX-1909/2020\|EPI_ISL_604225\|2020-0 | ···················· | ···················· | ····················· | ···················· | ···················· | ···················· |

| | | | | | | | | |
|---|---|---|---|---|---|---|---|---|
| N | 984 | hCoV-19/USA/MI-QDX-1943/2020\|EPI_ISL_604226\|2020-0 | ···················· | ···················· | ····················· | ···················· | ···················· | ···················· |
| N | 985 | hCoV-19/USA/MI-QDX-1960/2020\|EPI_ISL_604227\|2020-0 | ···················· | ···················· | ····················· | ···················· | ···················· | ···················· |
| N | 986 | hCoV-19/USA/NJ-QDX-1883/2020\|EPI_ISL_604228\|2020-0 | ···················· | ···················· | ····················· | ···················· | ···················· | ···················· |
| N | 987 | hCoV-19/USA/NJ-QDX-1886/2020\|EPI_ISL_604229\|2020-0 | ···················· | ···················· | ····················· | ···················· | ···················· | ···················· |
| N | 988 | hCoV-19/USA/NJ-QDX-1887/2020\|EPI_ISL_604230\|2020-0 | ···················· | ···················· | ····················· | ···················· | ···················· | ···················· |
| N | 989 | hCoV-19/USA/NJ-QDX-1900/2020\|EPI_ISL_604231\|2020-0 | ···················· | ···················· | ····················· | ···················· | ···················· | ···················· |
| N | 990 | hCoV-19/USA/NJ-QDX-1957/2020\|EPI_ISL_604232\|2020-0 | ···················· | ···················· | ····················· | ···················· | ···················· | ···················· |
| N | 991 | hCoV-19/USA/NJ-QDX-1968/2020\|EPI_ISL_604233\|2020-0 | ···················· | ···················· | ····················· | ···················· | ···················· | ···················· |
| N | 992 | hCoV-19/USA/NJ-QDX-1974/2020\|EPI_ISL_604234\|2020-0 | ···················· | ···················· | ····················· | ···················· | ···················· | ···················· |
| N | 993 | hCoV-19/USA/NY-QDX-1874/2020\|EPI_ISL_604235\|2020-0 | ···················· | ···················· | ····················· | ···················· | ···················· | ···················· |
| N | 994 | hCoV-19/USA/NY-QDX-1876/2020\|EPI_ISL_604236\|2020-0 | ···················· | ···················· | ····················· | ···················· | ···················· | ···················· |
| N | 995 | hCoV-19/USA/NY-QDX-1877/2020\|EPI_ISL_604237\|2020-0 | ···················· | ···················· | ····················· | ···················· | ···················· | ···················· |
| N | 996 | hCoV-19/USA/NY-QDX-1878/2020\|EPI_ISL_604238\|2020-0 | ···················· | ···················· | ····················· | ···················· | ···················· | ···················· |
| N | 997 | hCoV-19/USA/NY-QDX-1884/2020\|EPI_ISL_604239\|2020-0 | ···················· | ···················· | ····················· | ···················· | ···················· | ···················· |
| N | 998 | hCoV-19/USA/WI-UW-1620/2020\|EPI_ISL_605490\|2020-09 | ···················· | ···················· | ····················· | ···················· | ···················· | ···················· |
| N | 999 | hCoV-19/USA/WI-UW-1621/2020\|EPI_ISL_605491\|2020-09 | ···················· | ···················· | ····················· | ···················· | ···················· | ···················· |
| N | 1000 | hCoV-19/USA/WI-UW-1622/2020\|EPI_ISL_605492\|2020-09 | ···················· | ···················· | ····················· | ···················· | ···················· | ···················· |
| N | 1001 | hCoV-19/USA/WI-UW-1615/2020\|EPI_ISL_605493\|2020-09 | ···················· | ···················· | ····················· | ···················· | ···················· | ···················· |
| N | 1002 | hCoV-19/USA/WI-UW-1616/2020\|EPI_ISL_605494\|2020-09 | ···················· | ···················· | ····················· | ···················· | ···················· | ···················· |
| N | 1004 | hCoV-19/USA/WI-UW-1618/2020\|EPI_ISL_605496\|2020-09 | ···················· | ···················· | ····················· | ···················· | ···················· | ···················· |
| N | 1005 | hCoV-19/USA/IL-UW-1625/2020\|EPI_ISL_605499\|2020-09 | ···················· | ···················· | ····················· | ···················· | ···················· | ···················· |
| N | 1007 | hCoV-19/USA/WI-UW-1677/2020\|EPI_ISL_605551\|2020-09 | ···················· | ···················· | ····················· | ···················· | ···················· | ···················· |
| N | 1008 | hCoV-19/USA/WI-UW-1679/2020\|EPI_ISL_605552\|2020-09 | ···················· | ···················· | ····················· | ···················· | ···················· | ···················· |
| N | 1013 | hCoV-19/USA/WI-UW-1685/2020\|EPI_ISL_605558\|2020-09 | ···················· | ···················· | ····················· | ···················· | ···················· | ···················· |
| N | 1016 | hCoV-19/USA/WI-UW-1688/2020\|EPI_ISL_605561\|2020-09 | ···················· | ···················· | ····················· | ···················· | ···················· | ···················· |
| N | 1018 | hCoV-19/USA/WI-UW-1690/2020\|EPI_ISL_605563\|2020-09 | ···················· | ·············A······ | ····················· | ···················· | ···················· | ···················· |
| N | 1021 | hCoV-19/USA/WI-UW-1693/2020\|EPI_ISL_605566\|2020-09 | ···················· | ···················· | ····················· | ···················· | ···················· | ···················· |
| N | 1022 | hCoV-19/USA/WI-UW-1694/2020\|EPI_ISL_605567\|2020-09 | ···················· | ···················· | ····················· | ···················· | ···················· | ···················· |
| N | 1024 | hCoV-19/USA/WI-UW-1696/2020\|EPI_ISL_605569\|2020-09 | ···················· | ···················· | ····················· | ···················· | ···················· | ···················· |
| N | 1025 | hCoV-19/Australia/WA276/2020\|EPI_ISL_596819\|2020-1 | ···················· | ·A·················· | ····················· | ···················· | ···················· | ···················· |
| N | 1026 | hCoV-19/Australia/WA277/2020\|EPI_ISL_596820\|2020-0 | ···················· | ···················· | ····················· | ···················· | ···················· | ···················· |
| N | 1027 | hCoV-19/Australia/WA278/2020\|EPI_ISL_596821\|2020-1 | ···················· | ···················· | ····················· | ···················· | ···················· | ···················· |
| N | 1028 | hCoV-19/Australia/WA279/2020\|EPI_ISL_596822\|2020-0 | ···················· | ···················· | ····················· | ···················· | ···················· | ···················· |
| N | 1029 | hCoV-19/Australia/WA280/2020\|EPI_ISL_596823\|2020-0 | ···················· | ···················· | ····················· | ···················· | ···················· | ···················· |
| N | 1030 | hCoV-19/Australia/WA281/2020\|EPI_ISL_596824\|2020-0 | ···················· | ·············A······ | ····················· | ···················· | ···················· | ···················· |
| N | 1031 | hCoV-19/Australia/WA282/2020\|EPI_ISL_596825\|2020-0 | ···················· | ···················· | ····················· | ···················· | ···················· | ···················· |
| N | 1032 | hCoV-19/Australia/WA283/2020\|EPI_ISL_596826\|2020-0 | ···················· | ···················· | ····················· | ···················· | ···················· | ···················· |
| N | 1033 | hCoV-19/Australia/WA284/2020\|EPI_ISL_596827\|2020-0 | ···················· | ···················· | ····················· | ···················· | ···················· | ···················· |
| N | 1034 | hCoV-19/Australia/WA285/2020\|EPI_ISL_596828\|2020-0 | ···················· | ···················· | ····················· | ···················· | ···················· | ···················· |
| N | 1035 | hCoV-19/Australia/WA286/2020\|EPI_ISL_596829\|2020-0 | ···················· | ···················· | ····················· | ···················· | ···················· | ···················· |
| N | 1036 | hCoV-19/Australia/WA287/2020\|EPI_ISL_596830\|2020-0 | ···················· | ···················· | ····················· | ···················· | ···················· | ···················· |
| N | 1037 | hCoV-19/Australia/WA288/2020\|EPI_ISL_596831\|2020-0 | ···················· | ···················· | ····················· | ···················· | ···················· | ···················· |
| N | 1038 | hCoV-19/Australia/WA289/2020\|EPI_ISL_596832\|2020-0 | ···················· | ···················· | ····················· | ···················· | ···················· | ···················· |
| N | 1039 | hCoV-19/Australia/WA290/2020\|EPI_ISL_596833\|2020-0 | ···················· | ···················· | ····················· | ···················· | ···················· | ···················· |
| N | 1040 | hCoV-19/Australia/WA291/2020\|EPI_ISL_596834\|2020-0 | ···················· | ···················· | ····················· | ···················· | ···················· | ···················· |
| N | 1041 | hCoV-19/Australia/WA292/2020\|EPI_ISL_596835\|2020-0 | ···················· | ···················· | ····················· | ···················· | ···················· | ···················· |
| N | 1042 | hCoV-19/Australia/WA293/2020\|EPI_ISL_596836\|2020-0 | ···················· | ···················· | ····················· | ···················· | ···················· | ···················· |
| N | 1043 | hCoV-19/Australia/WA294/2020\|EPI_ISL_596837\|2020-0 | ···················· | ···················· | ····················· | ···················· | ···················· | ···················· |
| N | 1044 | hCoV-19/Australia/WA295/2020\|EPI_ISL_596838\|2020-0 | ···················· | ···················· | ····················· | ···················· | ···················· | ···················· |
| N | 1045 | hCoV-19/Australia/WA296/2020\|EPI_ISL_596839\|2020-0 | ···················· | ···················· | ····················· | ···················· | ···················· | ···················· |
| N | 1046 | hCoV-19/Australia/WA297/2020\|EPI_ISL_596840\|2020-0 | ···················· | ···················· | ····················· | ···················· | ···················· | ···················· |
| N | 1047 | hCoV-19/Australia/WA298/2020\|EPI_ISL_596841\|2020-0 | ···················· | ···················· | ····················· | ···················· | ···················· | ···················· |
| N | 1048 | hCoV-19/Australia/WA299/2020\|EPI_ISL_596842\|2020-1 | ···················· | ···················· | ····················· | ···················· | ···················· | ···················· |
| N | 1049 | hCoV-19/Australia/WA300/2020\|EPI_ISL_596843\|2020-0 | ···················· | ···················· | ····················· | ···················· | ···················· | ···················· |
| N | 1050 | hCoV-19/Australia/WA301/2020\|EPI_ISL_596844\|2020-0 | ···················· | ···················· | ····················· | ···················· | ···················· | ···················· |
| N | 1051 | hCoV-19/Australia/WA302/2020\|EPI_ISL_596845\|2020-0 | ···················· | ···················· | ····················· | ···················· | ···················· | ···················· |
| N | 1052 | hCoV-19/Australia/WA303/2020\|EPI_ISL_596846\|2020-0 | ···················· | ···················· | ····················· | ···················· | ···················· | ···················· |
| N | 1053 | hCoV-19/Australia/WA304/2020\|EPI_ISL_596847\|2020-0 | ···················· | ···················· | ····················· | ···················· | ···················· | ···················· |
| N | 1054 | hCoV-19/Australia/WA305/2020\|EPI_ISL_596848\|2020-0 | ···················· | ···················· | ····················· | ···················· | ···················· | ···················· |
| N | 1055 | hCoV-19/Australia/WA306/2020\|EPI_ISL_596849\|2020-0 | ···················· | ···················· | ····················· | ···················· | ···················· | ···················· |
| N | 1056 | hCoV-19/Australia/WA307/2020\|EPI_ISL_596850\|2020-0 | ···················· | ···················· | ····················· | ···················· | ···················· | ···················· |
| N | 1057 | hCoV-19/Australia/WA308/2020\|EPI_ISL_596851\|2020-0 | ···················· | ···················· | ····················· | ···················· | ···················· | ···················· |
| N | 1058 | hCoV-19/Australia/WA309/2020\|EPI_ISL_596852\|2020-0 | ···················· | ···················· | ····················· | ···················· | ···················· | ···················· |
| N | 1059 | hCoV-19/Australia/WA310/2020\|EPI_ISL_596853\|2020-0 | ···················· | ···················· | ····················· | ···················· | ···················· | ···················· |
| N | 1060 | hCoV-19/Australia/WA311/2020\|EPI_ISL_596854\|2020-0 | ···················· | ···················· | ····················· | ···················· | ···················· | ···················· |
| N | 1061 | hCoV-19/Australia/WA312/2020\|EPI_ISL_596855\|2020-0 | ···················· | ···················· | ····················· | ···················· | ···················· | ···················· |
| N | 1062 | hCoV-19/Australia/WA313/2020\|EPI_ISL_596856\|2020-0 | ···················· | ···················· | ····················· | ···················· | ···················· | ···················· |
| N | 1063 | hCoV-19/Australia/WA314/2020\|EPI_ISL_596857\|2020-0 | ···················· | ···················· | ····················· | ···················· | ···················· | ···················· |
| N | 1064 | hCoV-19/Australia/WA315/2020\|EPI_ISL_596858\|2020-0 | ···················· | ···················· | ····················· | ···················· | ···················· | ···················· |
| N | 1065 | hCoV-19/Australia/WA316/2020\|EPI_ISL_596859\|2020-0 | ···················· | ···················· | ····················· | ···················· | ···················· | ···················· |
| N | 1066 | hCoV-19/Australia/WA317/2020\|EPI_ISL_596860\|2020-0 | ···················· | ···················· | ····················· | ···················· | ···················· | ···················· |
| N | 1067 | hCoV-19/Australia/WA318/2020\|EPI_ISL_596861\|2020-0 | ···················· | ···················· | ····················· | ···················· | ···················· | ···················· |
| N | 1068 | hCoV-19/Scotland/QEUH-A21087/2020\|EPI_ISL_599000\|2 | ···················· | ···················· | ····················· | ···················· | ···················· | ···················· |
| N | 1069 | hCoV-19/Scotland/QEUH-A20D72/2020\|EPI_ISL_599001\|2 | ···················· | ···················· | ····················· | ···················· | ···················· | ···················· |
| N | 1070 | hCoV-19/Scotland/QEUH-A1FDD0/2020\|EPI_ISL_599002\|2 | ···················· | ···················· | ····················· | ···················· | ···················· | ···················· |
| N | 1071 | hCoV-19/Scotland/QEUH-A1F8B1/2020\|EPI_ISL_599003\|2 | ···················· | ···················· | ····················· | ···················· | ···················· | ···················· |
| N | 1072 | hCoV-19/Scotland/QEUH-A2253C/2020\|EPI_ISL_599004\|2 | ···················· | ···················· | ····················· | ···················· | ···················· | ···················· |
| N | 1073 | hCoV-19/Scotland/QEUH-A1FAF7/2020\|EPI_ISL_599005\|2 | ···················· | ···················· | ····················· | ···················· | ···················· | ···················· |
| N | 1074 | hCoV-19/Scotland/QEUH-A20DDC/2020\|EPI_ISL_599006\|2 | ···················· | ···················· | ····················· | ···················· | ···················· | ···················· |
| N | 1075 | hCoV-19/Scotland/QEUH-A1FB21/2020\|EPI_ISL_599007\|2 | ···················· | ···················· | ····················· | ···················· | ···················· | ···················· |

| | | | | | | | | |
|---|---|---|---|---|---|---|---|---|
| N | 1076 | hCoV-19/Scotland/QEUH-A1F7B4/2020\|EPI_ISL_599008\|2 | .................... | .................... | .................... | .................... | .................... | .................... |
| N | 1077 | hCoV-19/Scotland/QEUH-A1FBC7/2020\|EPI_ISL_599009\|2 | .................... | .................... | .................... | .................... | .................... | .................... |
| N | 1078 | hCoV-19/Scotland/QEUH-A22569/2020\|EPI_ISL_599011\|2 | .................... | .................... | .................... | .................... | .................... | .................... |
| N | 1079 | hCoV-19/Scotland/QEUH-A20440/2020\|EPI_ISL_599012\|2 | .................... | .................... | .................... | .................... | .................... | .................... |
| N | 1080 | hCoV-19/Scotland/QEUH-A1F63F/2020\|EPI_ISL_599013\|2 | .................... | .................... | .................... | .................... | .................... | .................... |
| N | 1081 | hCoV-19/Scotland/QEUH-A221B0/2020\|EPI_ISL_599014\|2 | .................... | .................... | .................... | .................... | .................... | .................... |
| N | 1082 | hCoV-19/Scotland/QEUH-A20941/2020\|EPI_ISL_599015\|2 | .................... | .................... | .................... | .................... | .................... | .................... |
| N | 1083 | hCoV-19/Scotland/QEUH-A1F796/2020\|EPI_ISL_599016\|2 | .................... | .................... | .................... | .................... | .................... | .................... |
| N | 1084 | hCoV-19/Scotland/QEUH-A1F523/2020\|EPI_ISL_599017\|2 | .................... | .................... | .................... | .................... | .................... | .................... |
| N | 1085 | hCoV-19/Scotland/QEUH-A1FBA9/2020\|EPI_ISL_599040\|2 | .................... | .................... | .................... | .................... | .................... | .................... |
| N | 1086 | hCoV-19/Scotland/QEUH-A1F611/2020\|EPI_ISL_599041\|2 | .................... | .................... | .................... | .................... | .................... | .................... |
| N | 1087 | hCoV-19/Scotland/QEUH-A226FD/2020\|EPI_ISL_599042\|2 | .................... | .................... | .................... | .................... | .................... | .................... |
| N | 1088 | hCoV-19/Scotland/QEUH-A1F699/2020\|EPI_ISL_599044\|2 | .................... | .................... | .................... | .................... | .................... | .................... |
| N | 1089 | hCoV-19/Scotland/QEUH-A1F909/2020\|EPI_ISL_599045\|2 | .................... | .................... | .................... | .................... | .................... | .................... |
| N | 1090 | hCoV-19/Scotland/QEUH-A1F73C/2020\|EPI_ISL_599046\|2 | .................... | .................... | .................... | .................... | .................... | .................... |
| N | 1091 | hCoV-19/Scotland/QEUH-A2104B/2020\|EPI_ISL_599047\|2 | .................... | .................... | .................... | .................... | .................... | .................... |
| N | 1092 | hCoV-19/Scotland/QEUH-A1FAE8/2020\|EPI_ISL_599048\|2 | .................... | .................... | .................... | .................... | .................... | .................... |
| N | 1093 | hCoV-19/Scotland/QEUH-A20E51/2020\|EPI_ISL_599060\|2 | .................... | .................... | .................... | .................... | .................... | .................... |
| N | 1094 | hCoV-19/Scotland/QEUH-A20413/2020\|EPI_ISL_599061\|2 | .................... | .................... | .................... | .................... | .................... | .................... |
| N | 1095 | hCoV-19/Scotland/QEUH-A1FBD6/2020\|EPI_ISL_599062\|2 | .................... | .................... | .................... | .................... | .................... | .................... |
| N | 1096 | hCoV-19/Scotland/QEUH-A20950/2020\|EPI_ISL_599063\|2 | .................... | .................... | .................... | .................... | .................... | .................... |
| N | 1097 | hCoV-19/Scotland/QEUH-A2251E/2020\|EPI_ISL_599064\|2 | .................... | .................... | .................... | .................... | .................... | .................... |
| N | 1098 | hCoV-19/Scotland/QEUH-A1FA33/2020\|EPI_ISL_599065\|2 | .................... | .................... | .................... | .................... | .................... | .................... |
| N | 1099 | hCoV-19/Scotland/QEUH-A1FC00/2020\|EPI_ISL_599066\|2 | .................... | .................... | .................... | .................... | .................... | .................... |
| N | 1100 | hCoV-19/Scotland/QEUH-A1F541/2020\|EPI_ISL_599067\|2 | .................... | .................... | .................... | .................... | .................... | .................... |
| N | 1101 | hCoV-19/Scotland/QEUH-A203E9/2020\|EPI_ISL_599068\|2 | .................... | .................... | .................... | .................... | .................... | .................... |
| N | 1102 | hCoV-19/Scotland/QEUH-A2103C/2020\|EPI_ISL_599069\|2 | .................... | .................... | .................... | .................... | .................... | .................... |
| N | 1103 | hCoV-19/Scotland/QEUH-A1FF9E/2020\|EPI_ISL_599070\|2 | .................... | .................... | .................... | .................... | .................... | .................... |
| N | 1104 | hCoV-19/Scotland/QEUH-A1F7D2/2020\|EPI_ISL_599071\|2 | .................... | .................... | .................... | .................... | .................... | .................... |
| N | 1105 | hCoV-19/Scotland/QEUH-A1F6C6/2020\|EPI_ISL_599072\|2 | .................... | .................... | .................... | .................... | .................... | .................... |
| N | 1106 | hCoV-19/Scotland/QEUH-A20CEE/2020\|EPI_ISL_599073\|2 | .................... | .................... | .................... | .................... | .................... | .................... |
| N | 1107 | hCoV-19/Scotland/QEUH-A1FACA/2020\|EPI_ISL_599074\|2 | .................... | .................... | .................... | .................... | .................... | .................... |
| N | 1108 | hCoV-19/Scotland/QEUH-A1FE82/2020\|EPI_ISL_599076\|2 | .................... | .................... | .................... | .................... | .................... | .................... |
| N | 1109 | hCoV-19/Scotland/QEUH-A1F787/2020\|EPI_ISL_599078\|2 | .................... | .................... | .................... | .................... | .................... | .................... |
| N | 1110 | hCoV-19/Scotland/QEUH-A1FFBC/2020\|EPI_ISL_599079\|2 | .................... | .................... | .................... | .................... | .................... | .................... |
| N | 1111 | hCoV-19/Scotland/QEUH-A20D36/2020\|EPI_ISL_599080\|2 | .................... | .................... | .................... | .................... | .................... | .................... |
| N | 1112 | hCoV-19/Scotland/QEUH-A20DBE/2020\|EPI_ISL_599081\|2 | .................... | .................... | .................... | .................... | .................... | .................... |
| N | 1113 | hCoV-19/Scotland/QEUH-A1FE73/2020\|EPI_ISL_599082\|2 | .................... | .................... | .................... | .................... | .................... | .................... |
| N | 1114 | hCoV-19/Scotland/QEUH-A20E15/2020\|EPI_ISL_599083\|2 | .................... | .................... | .................... | .................... | .................... | .................... |
| N | 1115 | hCoV-19/Scotland/QEUH-A1FF34/2020\|EPI_ISL_599084\|2 | .................... | .................... | .................... | .................... | .................... | .................... |
| N | 1116 | hCoV-19/Scotland/QEUH-A1FB5E/2020\|EPI_ISL_599085\|2 | .................... | .................... | .................... | .................... | .................... | .................... |
| N | 1117 | hCoV-19/Scotland/QEUH-A1F5D8/2020\|EPI_ISL_599086\|2 | .................... | .................... | .................... | .................... | .................... | .................... |
| N | 1118 | hCoV-19/Scotland/QEUH-A1F80C/2020\|EPI_ISL_599087\|2 | .................... | .................... | .................... | .................... | .................... | .................... |
| N | 1119 | hCoV-19/Scotland/QEUH-A1F75A/2020\|EPI_ISL_599088\|2 | .................... | .................... | .................... | .................... | .................... | .................... |
| N | 1120 | hCoV-19/Scotland/QEUH-A1F70F/2020\|EPI_ISL_599089\|2 | .................... | .................... | .................... | .................... | .................... | .................... |
| N | 1121 | hCoV-19/Scotland/QEUH-A20404/2020\|EPI_ISL_599090\|2 | .................... | .................... | .................... | .................... | .................... | .................... |
| N | 1122 | hCoV-19/Scotland/QEUH-A1F82A/2020\|EPI_ISL_599091\|2 | .................... | .................... | .................... | .................... | .................... | .................... |
| N | 1123 | hCoV-19/Scotland/QEUH-A1FDA3/2020\|EPI_ISL_599092\|2 | .................... | .................... | .................... | .................... | .................... | .................... |
| N | 1124 | hCoV-19/Scotland/QEUH-A20CFD/2020\|EPI_ISL_599093\|2 | .................... | .................... | .................... | .................... | .................... | .................... |
| N | 1125 | hCoV-19/Scotland/QEUH-A1FA8E/2020\|EPI_ISL_599094\|2 | .................... | .................... | .................... | .................... | .................... | .................... |
| N | 1126 | hCoV-19/Scotland/QEUH-A1FDEF/2020\|EPI_ISL_599095\|2 | .................... | .................... | .................... | .................... | .................... | .................... |
| N | 1127 | hCoV-19/Scotland/QEUH-A226C0/2020\|EPI_ISL_599096\|2 | .................... | .................... | .................... | .................... | .................... | .................... |
| N | 1128 | hCoV-19/Scotland/QEUH-A2254B/2020\|EPI_ISL_599097\|2 | .................... | .................... | .................... | .................... | .................... | .................... |
| N | 1129 | hCoV-19/Scotland/QEUH-A225B4/2020\|EPI_ISL_599098\|2 | .................... | .................... | .................... | .................... | .................... | .................... |
| N | 1130 | hCoV-19/Scotland/QEUH-A1F875/2020\|EPI_ISL_599099\|2 | .................... | .................... | .................... | .................... | .................... | .................... |
| N | 1131 | hCoV-19/Scotland/QEUH-9F6787/2020\|EPI_ISL_600639\|2 | .................... | .................... | .................... | .................... | .................... | .................... |
| N | 1132 | hCoV-19/Scotland/QEUH-9F5E0B/2020\|EPI_ISL_600640\|2 | .................... | .................... | .................... | .................... | .................... | .................... |
| N | 1133 | hCoV-19/Scotland/QEUH-9F4473/2020\|EPI_ISL_600641\|2 | .................... | .................... | .................... | .................... | .................... | .................... |
| N | 1134 | hCoV-19/Scotland/QEUH-9F6A7F/2020\|EPI_ISL_600642\|2 | .................... | .................... | .................... | .................... | .................... | .................... |
| N | 1135 | hCoV-19/Scotland/QEUH-9F6B21/2020\|EPI_ISL_600644\|2 | .................... | .................... | .................... | .................... | .................... | .................... |
| N | 1136 | hCoV-19/Scotland/QEUH-9F6B5E/2020\|EPI_ISL_600645\|2 | .................... | .................... | .................... | .................... | .................... | .................... |
| N | 1137 | hCoV-19/Scotland/QEUH-9F658D/2020\|EPI_ISL_600646\|2 | .................... | .................... | .................... | .................... | .................... | .................... |
| N | 1138 | hCoV-19/Scotland/QEUH-9F4446/2020\|EPI_ISL_600647\|2 | .................... | .................... | .................... | .................... | .................... | .................... |
| N | 1139 | hCoV-19/Scotland/QEUH-9F6A15/2020\|EPI_ISL_600648\|2 | .................... | .................... | .................... | .................... | .................... | .................... |
| N | 1140 | hCoV-19/Scotland/QEUH-9F43EF/2020\|EPI_ISL_600649\|2 | .................... | .................... | .................... | .................... | .................... | .................... |
| N | 1141 | hCoV-19/Scotland/QEUH-9F69AF/2020\|EPI_ISL_600650\|2 | .................... | .................... | .................... | .................... | .................... | .................... |
| N | 1142 | hCoV-19/Scotland/QEUH-9F6796/2020\|EPI_ISL_600651\|2 | .................... | .................... | .................... | .................... | .................... | .................... |
| N | 1143 | hCoV-19/Scotland/QEUH-9F66B7/2020\|EPI_ISL_600652\|2 | .................... | .................... | .................... | .................... | .................... | .................... |
| N | 1144 | hCoV-19/Scotland/QEUH-9F6523/2020\|EPI_ISL_600653\|2 | .................... | .................... | .................... | .................... | .................... | .................... |
| N | 1145 | hCoV-19/Scotland/QEUH-9F44FB/2020\|EPI_ISL_600654\|2 | .................... | .................... | .................... | .................... | .................... | .................... |
| N | 1146 | hCoV-19/Scotland/QEUH-9F67E1/2020\|EPI_ISL_600655\|2 | .................... | .................... | .................... | .................... | .................... | .................... |
| N | 1147 | hCoV-19/Scotland/QEUH-9F5F53/2020\|EPI_ISL_600656\|2 | .................... | .................... | .................... | .................... | .................... | .................... |
| N | 1148 | hCoV-19/Scotland/QEUH-9F4516/2020\|EPI_ISL_600657\|2 | .................... | .................... | .................... | .................... | .................... | .................... |
| N | 1149 | hCoV-19/Scotland/QEUH-9F6550/2020\|EPI_ISL_600658\|2 | .................... | .................... | .................... | .................... | .................... | .................... |
| N | 1150 | hCoV-19/Scotland/QEUH-9F6541/2020\|EPI_ISL_600659\|2 | .................... | .................... | .................... | .................... | .................... | .................... |
| N | 1151 | hCoV-19/Scotland/QEUH-9F4C99/2020\|EPI_ISL_600660\|2 | .................... | .................... | .................... | .................... | .................... | .................... |
| N | 1152 | hCoV-19/Scotland/QEUH-9F43FE/2020\|EPI_ISL_600661\|2 | .................... | .................... | .................... | .................... | .................... | .................... |
| N | 1153 | hCoV-19/Scotland/QEUH-9F675A/2020\|EPI_ISL_600662\|2 | .................... | .................... | .................... | .................... | .................... | .................... |
| N | 1154 | hCoV-19/Scotland/QEUH-9F66F3/2020\|EPI_ISL_600663\|2 | .................... | .................... | .................... | .................... | .................... | .................... |
| N | 1155 | hCoV-19/Scotland/QEUH-9F66C6/2020\|EPI_ISL_600664\|2 | .................... | .................... | .................... | .................... | .................... | .................... |

N 1156 hCoV-19/Scotland/QEUH-9F66D5/2020|EPI_ISL_600665|2 ···················· ···················· ···················· ···················· ···················· ····················
N 1157 hCoV-19/Scotland/QEUH-9F67C3/2020|EPI_ISL_600666|2 ···················· ···················· ···················· ···················· ···················· ····················
N 1158 hCoV-19/Scotland/QEUH-9F6981/2020|EPI_ISL_600667|2 ···················· ···················· ···················· ···················· ···················· ····················
N 1159 hCoV-19/Scotland/QEUH-9F6927/2020|EPI_ISL_600668|2 ···················· ···················· ···················· ···················· ···················· ····················
N 1160 hCoV-19/Scotland/QEUH-9F65AB/2020|EPI_ISL_600669|2 ···················· ···················· ···················· ···················· ···················· ····················
N 1161 hCoV-19/Scotland/QEUH-9F6B30/2020|EPI_ISL_600670|2 ···················· ···················· ···················· ···················· ···················· ····················
N 1162 hCoV-19/Scotland/QEUH-9F674B/2020|EPI_ISL_600671|2 ···················· ···················· ···················· ···················· ···················· ····················
N 1163 hCoV-19/Scotland/QEUH-9F467D/2020|EPI_ISL_600672|2 ···················· ···················· ···················· ···················· ···················· ····················
N 1164 hCoV-19/Scotland/QEUH-9F5D77/2020|EPI_ISL_600673|2 ···················· ···················· ···················· ···················· ···················· ····················
N 1165 hCoV-19/Scotland/QEUH-9F4701/2020|EPI_ISL_600674|2 ···················· ···················· ···················· ···················· ···················· ····················
N 1166 hCoV-19/Scotland/QEUH-9F45F8/2020|EPI_ISL_600675|2 ···················· ···················· ···················· ···················· ···················· ····················
N 1167 hCoV-19/Scotland/QEUH-9F6A06/2020|EPI_ISL_600676|2 ···················· ·A·················· ···················· ···················· ···················· ··A·················
N 1168 hCoV-19/Scotland/QEUH-9F46E6/2020|EPI_ISL_600677|2 ···················· ···················· ···················· ···················· ···················· ····················
N 1169 hCoV-19/Scotland/QEUH-9F6A51/2020|EPI_ISL_600678|2 ···················· ···················· ···················· ···················· ···················· ····················
N 1170 hCoV-19/Scotland/QEUH-9CF787/2020|EPI_ISL_601970|2 ···················· ···················· ···················· ···················· ···················· ····················
N 1171 hCoV-19/Scotland/QEUH-9CE339/2020|EPI_ISL_601972|2 ···················· ···················· ···················· ···················· ···················· ····················
N 1172 hCoV-19/Scotland/QEUH-9CEB13/2020|EPI_ISL_601974|2 ···················· ···················· ···················· ···················· ···················· ····················
N 1173 hCoV-19/Scotland/QEUH-9CF4DB/2020|EPI_ISL_601975|2 ···················· ···················· ···················· ···················· ···················· ····················
N 1174 hCoV-19/Scotland/QEUH-9CE481/2020|EPI_ISL_601976|2 ···················· ···················· ···················· ···················· ······T············· ····················
N 1175 hCoV-19/Scotland/QEUH-9CE348/2020|EPI_ISL_601978|2 ···················· ···················· ···················· ···················· ···················· ····················
N 1176 hCoV-19/Scotland/QEUH-9CF013/2020|EPI_ISL_601979|2 ···················· ···················· ···················· ···················· ···················· ····················
N 1177 hCoV-19/Scotland/QEUH-9CF611/2020|EPI_ISL_601980|2 ···················· ···················· ···················· ···················· ···················· ····················
N 1178 hCoV-19/Scotland/QEUH-9CE427/2020|EPI_ISL_601981|2 ···················· ···················· ···················· ···················· ···················· ····················
N 1179 hCoV-19/Scotland/QEUH-9CE9A0/2020|EPI_ISL_601982|2 ···················· ···················· ···················· ···················· ···················· ····················
N 1180 hCoV-19/Scotland/QEUH-9CEF53/2020|EPI_ISL_601983|2 ···················· ···················· ···················· ···················· ···················· ····················
N 1181 hCoV-19/Scotland/QEUH-9CE8EF/2020|EPI_ISL_601984|2 ···················· ···················· ···················· ···················· ···················· ····················
N 1182 hCoV-19/Scotland/QEUH-9CE946/2020|EPI_ISL_601985|2 ···················· ···················· ···················· ···················· ···················· ····················
N 1183 hCoV-19/Scotland/QEUH-9CEA25/2020|EPI_ISL_601986|2 ···················· ···················· ···················· ···················· ···················· ····················
N 1184 hCoV-19/Scotland/QEUH-9CE3FD/2020|EPI_ISL_601987|2 ···················· ···················· ···················· ···················· ···················· ····················
N 1185 hCoV-19/Scotland/QEUH-9CF329/2020|EPI_ISL_601988|2 ···················· ···················· ···················· ···················· ······T············· ····················
N 1186 hCoV-19/Scotland/QEUH-9CF67B/2020|EPI_ISL_601989|2 ···················· ···················· ···················· ···················· ···················· ····················
N 1187 hCoV-19/Scotland/QEUH-9CF541/2020|EPI_ISL_601990|2 ···················· ···················· ···················· ···················· ···················· ····················
N 1188 hCoV-19/Scotland/QEUH-9CEADA/2020|EPI_ISL_601991|2 ···················· ···················· ···················· ···················· ···················· ····················
N 1189 hCoV-19/Scotland/QEUH-9CF64E/2020|EPI_ISL_601992|2 ···················· ···················· ···················· ···················· ···················· ····················
N 1190 hCoV-19/Scotland/QEUH-9CE973/2020|EPI_ISL_601994|2 ···················· ···················· ···················· ···················· ···················· ····················
N 1191 hCoV-19/Scotland/QEUH-9CE506/2020|EPI_ISL_601995|2 ···················· ···················· ···················· ···················· ···················· ····················
N 1192 hCoV-19/Scotland/QEUH-9CF80C/2020|EPI_ISL_601997|2 ···················· ···················· ···················· ···················· ···················· ····················
N 1193 hCoV-19/Scotland/QEUH-9CEC6B/2020|EPI_ISL_601998|2 ···················· ···················· ···················· ···················· ···················· ····················
N 1194 hCoV-19/Scotland/QEUH-9CF2D1/2020|EPI_ISL_601999|2 ···················· ···················· ···················· ···················· ···················· ····················
N 1195 hCoV-19/USA/UT-QDX-1789/2020|EPI_ISL_604121|2020-0 ···················· ···················· ···················· ···················· ···················· ····················
N 1196 hCoV-19/USA/UT-QDX-1790/2020|EPI_ISL_604122|2020-0 ···················· ···················· ···················· ···················· ···················· ····················
N 1197 hCoV-19/USA/UT-QDX-1793/2020|EPI_ISL_604123|2020-0 ···················· ···················· ···················· ···················· ···················· ····················
N 1198 hCoV-19/USA/UT-QDX-1794/2020|EPI_ISL_604124|2020-0 ···················· ···················· ···················· ···················· ···················· ····················
N 1199 hCoV-19/USA/UT-QDX-1795/2020|EPI_ISL_604125|2020-0 ···················· ···················· ···················· ···················· ···················· ····················
N 1200 hCoV-19/USA/UT-QDX-1796/2020|EPI_ISL_604126|2020-0 ···················· ···················· ···················· ···················· ···················· ····················
N 1201 hCoV-19/USA/UT-QDX-1797/2020|EPI_ISL_604127|2020-0 ···················· ···················· ···················· ···················· ···················· ····················
N 1202 hCoV-19/USA/UT-QDX-1799/2020|EPI_ISL_604128|2020-0 ···················· ···················· ···················· ···················· ···················· ····················
N 1203 hCoV-19/USA/UT-QDX-1800/2020|EPI_ISL_604129|2020-0 ···················· ···················· ···················· ···················· ···················· ····················
N 1204 hCoV-19/USA/UT-QDX-1801/2020|EPI_ISL_604130|2020-0 ···················· ···················· ···················· ···················· ···················· ····················
N 1205 hCoV-19/USA/UT-QDX-1804/2020|EPI_ISL_604131|2020-0 ···················· ···················· ···················· ···················· ···················· ····················
N 1206 hCoV-19/USA/UT-QDX-1805/2020|EPI_ISL_604132|2020-0 ···················· ···················· ···················· ···················· ···················· ····················
N 1207 hCoV-19/USA/UT-QDX-1806/2020|EPI_ISL_604133|2020-0 ···················· ···················· ···················· ···················· ···················· ····················
N 1208 hCoV-19/USA/UT-QDX-1807/2020|EPI_ISL_604134|2020-0 ···················· ···················· ···················· ···················· ···················· ····················
N 1209 hCoV-19/USA/UT-QDX-1808/2020|EPI_ISL_604135|2020-0 ···················· ···················· ···················· ···················· ···················· ····················
N 1210 hCoV-19/USA/UT-QDX-1809/2020|EPI_ISL_604136|2020-0 ···················· ···················· ···················· ···················· ···················· ····················
N 1211 hCoV-19/USA/UT-QDX-1811/2020|EPI_ISL_604137|2020-0 ···················· ···················· ···················· ···················· ···················· ····················
N 1212 hCoV-19/USA/UT-QDX-1812/2020|EPI_ISL_604138|2020-0 ···················· ···················· ···················· ···················· ···················· ····················
N 1213 hCoV-19/USA/UT-QDX-1815/2020|EPI_ISL_604139|2020-0 ···················· ···················· ···················· ···················· ···················· ····················
N 1214 hCoV-19/USA/UT-QDX-1816/2020|EPI_ISL_604140|2020-0 ···················· ···················· ···················· ···················· ···················· ····················
N 1215 hCoV-19/USA/UT-QDX-1818/2020|EPI_ISL_604141|2020-0 ···················· ···················· ···················· ···················· ···················· ····················
N 1216 hCoV-19/USA/UT-QDX-1822/2020|EPI_ISL_604142|2020-0 ···················· ···················· ···················· ···················· ···················· ····················
N 1217 hCoV-19/USA/UT-QDX-1823/2020|EPI_ISL_604143|2020-0 ···················· ···················· ···················· ···················· ···················· ····················
N 1218 hCoV-19/USA/UT-QDX-1825/2020|EPI_ISL_604144|2020-0 ···················· ···················· ···················· ···················· ···················· ····················
N 1219 hCoV-19/USA/UT-QDX-1827/2020|EPI_ISL_604145|2020-0 ···················· ···················· ···················· ···················· ···················· ····················
N 1220 hCoV-19/USA/UT-QDX-1828/2020|EPI_ISL_604146|2020-0 ···················· ···················· ···················· ···················· ···················· ····················
N 1221 hCoV-19/USA/UT-QDX-1829/2020|EPI_ISL_604147|2020-0 ···················· ···················· ···················· ···················· ···················· ····················
N 1222 hCoV-19/USA/UT-QDX-1830/2020|EPI_ISL_604148|2020-0 ···················· ···················· ···················· ···················· ···················· ····················
N 1223 hCoV-19/USA/UT-QDX-1831/2020|EPI_ISL_604149|2020-0 ···················· ···················· ···················· ···················· ···················· ····················
N 1224 hCoV-19/USA/UT-QDX-1832/2020|EPI_ISL_604150|2020-0 ···················· ···················· ···················· ···················· ···················· ····················
N 1225 hCoV-19/USA/UT-QDX-1833/2020|EPI_ISL_604151|2020-0 ···················· ···················· ···················· ···················· ···················· ····················
N 1226 hCoV-19/USA/UT-QDX-1834/2020|EPI_ISL_604152|2020-0 ···················· ···················· ···················· ···················· ···················· ····················
N 1227 hCoV-19/USA/UT-QDX-1835/2020|EPI_ISL_604153|2020-0 ···················· ···················· ···················· ···················· ···················· ····················
N 1228 hCoV-19/USA/UT-QDX-1836/2020|EPI_ISL_604154|2020-0 ···················· ···················· ···················· ···················· ···················· ····················
N 1229 hCoV-19/USA/UT-QDX-1838/2020|EPI_ISL_604155|2020-0 ···················· ···················· ···················· ···················· ···················· ····················
N 1230 hCoV-19/USA/UT-QDX-1839/2020|EPI_ISL_604156|2020-0 ···················· ···················· ···················· ···················· ···················· ····················
N 1231 hCoV-19/USA/UT-QDX-1841/2020|EPI_ISL_604157|2020-0 ···················· ···················· ···················· ···················· ···················· ····················
N 1232 hCoV-19/USA/UT-QDX-1842/2020|EPI_ISL_604158|2020-0 ···················· ···················· ···················· ···················· ···················· ····················
N 1233 hCoV-19/USA/UT-QDX-1843/2020|EPI_ISL_604159|2020-0 ···················· ···················· ···················· ···················· ···················· ····················
N 1234 hCoV-19/USA/UT-UPHL-2009728/2020|EPI_ISL_605400|20 ···················· ···················· ···················· ···················· ···················· ····················
N 1235 hCoV-19/USA/UT-UPHL-2009730/2020|EPI_ISL_605402|20 ···················· ···················· ···················· ···················· ···················· ····················

| | | | | | | | |
|---|---|---|---|---|---|---|---|
| N | 1236 hCoV-19/USA/UT-UPHL-2009731/2020\|EPI_ISL_605403\|20 | .................... | .................... | ..................... | .................... | .................... | .................... |
| N | 1237 hCoV-19/USA/WI-UW-1527/2020\|EPI_ISL_605405\|2020-09 | .................... | .................... | ..................... | .................... | .................... | .................... |
| N | 1238 hCoV-19/USA/WI-UW-1528/2020\|EPI_ISL_605406\|2020-09 | .................... | .................... | ..................... | .................... | .................... | .................... |
| N | 1240 hCoV-19/USA/WI-UW-1530/2020\|EPI_ISL_605408\|2020-09 | .................... | .................... | ..................... | .................... | .................... | .................... |
| N | 1241 hCoV-19/USA/WI-UW-1531/2020\|EPI_ISL_605409\|2020-09 | .................... | .................... | ..................... | .................... | .................... | .................... |
| N | 1242 hCoV-19/USA/WI-UW-1532/2020\|EPI_ISL_605410\|2020-09 | .................... | .................... | ..................... | .................... | .................... | .................... |
| N | 1243 hCoV-19/USA/WI-UW-1533/2020\|EPI_ISL_605411\|2020-09 | .................... | .................... | ..................... | .................... | .................... | .................... |
| N | 1244 hCoV-19/USA/WI-UW-1534/2020\|EPI_ISL_605412\|2020-09 | .................... | .................... | ..................... | .................... | .................... | .................... |
| N | 1247 hCoV-19/USA/WI-UW-1537/2020\|EPI_ISL_605415\|2020-09 | .................... | .................... | ..................... | .................... | .................... | .................... |
| N | 1248 hCoV-19/USA/WI-UW-1539/2020\|EPI_ISL_605417\|2020-09 | .................... | .................... | ..................... | .................... | .................... | .................... |
| N | 1249 hCoV-19/USA/WI-UW-1540/2020\|EPI_ISL_605418\|2020-09 | .................... | .................... | ..................... | .................... | .................... | .................... |
| N | 1250 hCoV-19/USA/WI-UW-1541/2020\|EPI_ISL_605419\|2020-09 | .................... | .................... | ..................... | .................... | .................... | .................... |
| N | 1261 hCoV-19/USA/WI-UW-1589/2020\|EPI_ISL_605466\|2020-09 | .................... | .................... | ..................... | .................... | .................... | .................... |
| N | 1262 hCoV-19/USA/WI-UW-1590/2020\|EPI_ISL_605467\|2020-09 | .................... | .................... | ..................... | .................... | .................... | .................... |
| N | 1263 hCoV-19/USA/WI-UW-1591/2020\|EPI_ISL_605468\|2020-09 | .................... | .................... | ..................... | .................... | .................... | .................... |
| N | 1264 hCoV-19/USA/WI-UW-1592/2020\|EPI_ISL_605469\|2020-09 | .................... | .................... | ..................... | .................... | .................... | .................... |
| N | 1267 hCoV-19/USA/WI-UW-1597/2020\|EPI_ISL_605474\|2020-09 | .................... | .................... | ..................... | .................... | .................... | .................... |
| N | 1268 hCoV-19/USA/WI-UW-1598/2020\|EPI_ISL_605475\|2020-09 | .................... | .................... | ..................... | .................... | .................... | .................... |
| N | 1269 hCoV-19/USA/WI-UW-1599/2020\|EPI_ISL_605476\|2020-09 | .................... | .................... | ..................... | .................... | .................... | .................... |
| N | 1270 hCoV-19/USA/WI-UW-1600/2020\|EPI_ISL_605477\|2020-09 | .................... | .................... | ..................... | .................... | .................... | .................... |
| N | 1272 hCoV-19/USA/WI-UW-1602/2020\|EPI_ISL_605479\|2020-09 | .................... | .................... | ..................... | .................... | .................... | .................... |
| N | 1273 hCoV-19/USA/WI-UW-1603/2020\|EPI_ISL_605480\|2020-09 | .................... | .................... | ..................... | .................... | .................... | .................... |
| N | 1275 hCoV-19/USA/WI-UW-1605/2020\|EPI_ISL_605482\|2020-09 | .................... | .................... | ..................... | .................... | .................... | .................... |
| N | 1276 hCoV-19/USA/WI-UW-1606/2020\|EPI_ISL_605483\|2020-09 | .................... | .................... | ..................... | .................... | .................... | .................... |
| N | 1277 hCoV-19/USA/WI-UW-1608/2020\|EPI_ISL_605485\|2020-09 | .................... | .................... | ..................... | .................... | .................... | .................... |
| N | 1278 hCoV-19/USA/WI-UW-1609/2020\|EPI_ISL_605486\|2020-09 | .................... | .................... | ..................... | .................... | .................... | .................... |
| N | 1279 hCoV-19/USA/WI-UW-1610/2020\|EPI_ISL_605487\|2020-09 | .................... | .................... | ..................... | .................... | .................... | .................... |
| N | 1280 hCoV-19/USA/WI-UW-1619/2020\|EPI_ISL_605489\|2020-09 | .................... | .................... | ..................... | .................... | .................... | .................... |
| N | 1281 hCoV-19/Australia/WA319/2020\|EPI_ISL_596862\|2020-0 | .................... | .................... | ..................... | .................... | .................... | .................... |
| N | 1282 hCoV-19/Ireland/OY-NVRL-73IRL45403/2020\|EPI_ISL_59 | .................... | .................... | ..................... | .................... | .................... | .................... |
| N | 1283 hCoV-19/Ireland/KE-NVRL-73IRL55350/2020\|EPI_ISL_59 | .................... | .................... | ..................... | .................... | .................... | .................... |
| N | 1284 hCoV-19/Ireland/LK-NVRL-73IRL35422/2020\|EPI_ISL_59 | .................... | .A.................. | ..................... | .................... | .................... | ..A................. |
| N | 1285 hCoV-19/Ireland/D-NVRL-73IRL62905/2020\|EPI_ISL_596 | .................... | .................... | ..................... | .................... | .................... | .................... |
| N | 1286 hCoV-19/Ireland/D-NVRL-73IRL38401/2020\|EPI_ISL_596 | .................... | .................... | ..................... | .................... | .................... | .................... |
| N | 1287 hCoV-19/Ireland/KE-NVRL-73IRL48809/2020\|EPI_ISL_59 | .................... | .................... | ..................... | .................... | .................... | .................... |
| N | 1288 hCoV-19/Ireland/OY-NVRL-73IRL44171/2020\|EPI_ISL_59 | .................... | .................... | ..................... | .................... | .................... | .................... |
| N | 1289 hCoV-19/Ireland/D-NVRL-73IRL76160/2020\|EPI_ISL_596 | .................... | .................... | ..................... | .................... | .................... | .................... |
| N | 1290 hCoV-19/Ireland/WX-NVRL-73IRL52567/2020\|EPI_ISL_59 | .................... | .................... | ..................... | .................... | .................... | .................... |
| N | 1291 hCoV-19/Ireland/D-NVRL-73IRL76164/2020\|EPI_ISL_596 | .................... | .................... | ..................... | .................... | .................... | .................... |
| N | 1292 hCoV-19/Scotland/QEUH-A1F8C0/2020\|EPI_ISL_599018\|2 | .................... | .................... | ..................... | .................... | .................... | .................... |
| N | 1293 hCoV-19/Scotland/QEUH-A22596/2020\|EPI_ISL_599019\|2 | .................... | .................... | ..................... | .................... | .................... | .................... |
| N | 1294 hCoV-19/Scotland/QEUH-A1F884/2020\|EPI_ISL_599020\|2 | .................... | .................... | ..................... | .................... | .................... | .................... |
| N | 1295 hCoV-19/Scotland/QEUH-A1FB6D/2020\|EPI_ISL_599021\|2 | .................... | .................... | ..................... | .................... | .................... | .................... |
| N | 1296 hCoV-19/Scotland/QEUH-A20E60/2020\|EPI_ISL_599022\|2 | .................... | .................... | ..................... | .................... | .................... | .................... |
| N | 1297 hCoV-19/Scotland/QEUH-A1F7F0/2020\|EPI_ISL_599023\|2 | .................... | .................... | ..................... | .................... | .................... | .................... |
| N | 1298 hCoV-19/Scotland/QEUH-A1FC2E/2020\|EPI_ISL_599024\|2 | .................... | .................... | ..................... | .................... | .................... | .................... |
| N | 1299 hCoV-19/Scotland/QEUH-A1F7A5/2020\|EPI_ISL_599025\|2 | .................... | .................... | ..................... | .................... | .................... | .................... |
| N | 1300 hCoV-19/Scotland/QEUH-A203CB/2020\|EPI_ISL_599026\|2 | .................... | .................... | ..................... | .................... | .................... | .................... |
| N | 1301 hCoV-19/Scotland/QEUH-A20D18/2020\|EPI_ISL_599027\|2 | .................... | .................... | ..................... | .................... | .................... | .................... |
| N | 1302 hCoV-19/Scotland/QEUH-A1F6B7/2020\|EPI_ISL_599028\|2 | .................... | .................... | ..................... | .................... | .................... | .................... |
| N | 1303 hCoV-19/Scotland/QEUH-A226B1/2020\|EPI_ISL_599029\|2 | .................... | .................... | ..................... | .................... | .................... | .................... |
| N | 1304 hCoV-19/Scotland/QEUH-A20D45/2020\|EPI_ISL_599030\|2 | .................... | .................... | ..................... | .................... | .................... | .................... |
| N | 1305 hCoV-19/Scotland/QEUH-A1FDFE/2020\|EPI_ISL_599031\|2 | .................... | .................... | ..................... | .................... | .................... | .................... |
| N | 1306 hCoV-19/Scotland/QEUH-A1F6D5/2020\|EPI_ISL_599032\|2 | .................... | .................... | ..................... | .................... | .................... | .................... |
| N | 1307 hCoV-19/Scotland/QEUH-A20E42/2020\|EPI_ISL_599033\|2 | .................... | .................... | ..................... | .................... | .................... | .................... |
| N | 1308 hCoV-19/Scotland/QEUH-A1FDC1/2020\|EPI_ISL_599034\|2 | .................... | .................... | ..................... | .................... | .................... | .................... |
| N | 1309 hCoV-19/Scotland/QEUH-A1FB9A/2020\|EPI_ISL_599035\|2 | .................... | .................... | ..................... | .................... | .................... | .................... |
| N | 1310 hCoV-19/Scotland/QEUH-A1F58D/2020\|EPI_ISL_599036\|2 | .................... | .................... | ..................... | .................... | .................... | .................... |
| N | 1311 hCoV-19/Scotland/QEUH-A1FBE5/2020\|EPI_ISL_599037\|2 | .................... | .................... | ..................... | .................... | .................... | .................... |
| N | 1312 hCoV-19/Scotland/QEUH-A225C3/2020\|EPI_ISL_599038\|2 | .................... | .................... | ..................... | .................... | .................... | .................... |
| N | 1313 hCoV-19/Scotland/QEUH-A1F66C/2020\|EPI_ISL_599039\|2 | .................... | .................... | ..................... | .................... | .................... | .................... |
| N | 1314 hCoV-19/Scotland/QEUH-A2255A/2020\|EPI_ISL_599050\|2 | .................... | .................... | ..................... | .................... | .................... | .................... |
| N | 1315 hCoV-19/Wales/QEUH-A203AD/2020\|EPI_ISL_599051\|2020 | .................... | .................... | ..................... | .................... | .................... | .................... |
| N | 1316 hCoV-19/Scotland/QEUH-A226EE/2020\|EPI_ISL_599052\|2 | .................... | .................... | ..................... | .................... | .................... | .................... |
| N | 1317 hCoV-19/Scotland/QEUH-A1FF70/2020\|EPI_ISL_599053\|2 | .................... | .................... | ..................... | .................... | .................... | .................... |
| N | 1318 hCoV-19/Scotland/QEUH-A20D90/2020\|EPI_ISL_599054\|2 | .................... | .................... | ..................... | .................... | .................... | .................... |
| N | 1319 hCoV-19/Scotland/QEUH-A1FB8B/2020\|EPI_ISL_599055\|2 | .................... | .................... | ..................... | .................... | .................... | .................... |
| N | 1320 hCoV-19/Scotland/QEUH-A1FC5B/2020\|EPI_ISL_599056\|2 | .................... | .................... | ..................... | .................... | .................... | .................... |
| N | 1321 hCoV-19/Scotland/QEUH-A1F68A/2020\|EPI_ISL_599057\|2 | .................... | .................... | ..................... | .................... | .................... | .................... |
| N | 1322 hCoV-19/Scotland/QEUH-A1FECE/2020\|EPI_ISL_599058\|2 | .................... | .................... | ..................... | .................... | .................... | .................... |
| N | 1323 hCoV-19/Scotland/QEUH-A203BC/2020\|EPI_ISL_599059\|2 | .................... | .................... | ..................... | .................... | .................... | .................... |
| N | 1324 hCoV-19/England/QEUH-9FFDF5/2020\|EPI_ISL_600501\|20 | .................... | .................... | ..................... | .................... | .................... | .................... |
| N | 1325 hCoV-19/England/QEUH-9FF979/2020\|EPI_ISL_600503\|20 | .................... | .................... | ..................... | .................... | .................... | .................... |
| N | 1326 hCoV-19/England/QEUH-9FF50C/2020\|EPI_ISL_600504\|20 | .................... | .................... | ..................... | .................... | .....T.............. | .................... |
| N | 1327 hCoV-19/England/QEUH-A00446/2020\|EPI_ISL_600506\|20 | .................... | .................... | ..................... | .................... | .................... | .................... |
| N | 1328 hCoV-19/England/QEUH-A00938/2020\|EPI_ISL_600509\|20 | .................... | .................... | ..................... | .................... | .................... | .................... |
| N | 1329 hCoV-19/Scotland/QEUH-9FF7AC/2020\|EPI_ISL_600511\|2 | .................... | .................... | ..................... | .................... | .................... | .................... |
| N | 1330 hCoV-19/England/QEUH-A00877/2020\|EPI_ISL_600513\|20 | .................... | .................... | ..................... | .................... | .................... | .................... |
| N | 1331 hCoV-19/Scotland/QEUH-9FB62B/2020\|EPI_ISL_600558\|2 | .................... | .................... | ..................... | .................... | .................... | .................... |
| N | 1332 hCoV-19/Scotland/QEUH-9FB1FD/2020\|EPI_ISL_600560\|2 | .................... | .................... | ..................... | .................... | .................... | .................... |

| | | | | | | | | |
|---|---|---|---|---|---|---|---|---|
| N | 1333 | hCoV-19/Scotland/QEUH-9FB44F/2020\|EPI_ISL_600561\|2 | .................... | .................... | .................... | .................... | .................... | .................... |
| N | 1334 | hCoV-19/Scotland/QEUH-9FB676/2020\|EPI_ISL_600563\|2 | .................... | .................... | .................... | .................... | .................... | .................... |
| N | 1335 | hCoV-19/Wales/QEUH-9FACFD/2020\|EPI_ISL_600565\|2020 | .................... | .................... | .................... | .................... | .................... | .................... |
| N | 1336 | hCoV-19/Scotland/QEUH-9FA3AD/2020\|EPI_ISL_600566\|2 | .................... | .................... | .................... | .................... | .................... | .................... |
| N | 1337 | hCoV-19/Scotland/QEUH-9FB39D/2020\|EPI_ISL_600568\|2 | .................... | .................... | .................... | .................... | .................... | .................... |
| N | 1338 | hCoV-19/Scotland/QEUH-9FB236/2020\|EPI_ISL_600569\|2 | .................... | .................... | .................... | .................... | .................... | .................... |
| N | 1339 | hCoV-19/Scotland/QEUH-9FA914/2020\|EPI_ISL_600571\|2 | .................... | .................... | .................... | .................... | .................... | .................... |
| N | 1340 | hCoV-19/Scotland/QEUH-9FB430/2020\|EPI_ISL_600572\|2 | .................... | .................... | .................... | .................... | .................... | .................... |
| N | 1341 | hCoV-19/Scotland/QEUH-9FA9C9/2020\|EPI_ISL_600573\|2 | .................... | .................... | .................... | .................... | .................... | .................... |
| N | 1342 | hCoV-19/Scotland/QEUH-9FB46D/2020\|EPI_ISL_600574\|2 | .................... | .................... | .................... | .................... | .................... | .................... |
| N | 1343 | hCoV-19/Scotland/QEUH-9FA97E/2020\|EPI_ISL_600575\|2 | .................... | .................... | .................... | .................... | .................... | .................... |
| N | 1344 | hCoV-19/Scotland/QEUH-9FB412/2020\|EPI_ISL_600576\|2 | .................... | .................... | .................... | .................... | .................... | .................... |
| N | 1345 | hCoV-19/Scotland/QEUH-9FA9D8/2020\|EPI_ISL_600577\|2 | .................... | .................... | .................... | .................... | .................... | .................... |
| N | 1346 | hCoV-19/Scotland/QEUH-9FB38E/2020\|EPI_ISL_600578\|2 | .................... | .................... | .................... | .................... | .................... | .................... |
| N | 1347 | hCoV-19/Scotland/QEUH-9FB1A2/2020\|EPI_ISL_600579\|2 | .................... | .................... | .................... | .................... | .................... | .................... |
| N | 1348 | hCoV-19/Scotland/QEUH-9FB3E8/2020\|EPI_ISL_600580\|2 | .................... | .................... | .................... | .................... | .................... | .................... |
| N | 1349 | hCoV-19/Scotland/QEUH-9FB47C/2020\|EPI_ISL_600581\|2 | .................... | .................... | .................... | .................... | .................... | .................... |
| N | 1350 | hCoV-19/Scotland/QEUH-9FB218/2020\|EPI_ISL_600582\|2 | .................... | .................... | .................... | .................... | .................... | .................... |
| N | 1351 | hCoV-19/Scotland/QEUH-9FB5A6/2020\|EPI_ISL_600583\|2 | .................... | .................... | .................... | .................... | .................... | .................... |
| N | 1352 | hCoV-19/Scotland/QEUH-9FB5F1/2020\|EPI_ISL_600584\|2 | .................... | .................... | .................... | .................... | .................... | .................... |
| N | 1353 | hCoV-19/Scotland/QEUH-9FB61C/2020\|EPI_ISL_600585\|2 | .................... | .................... | .................... | .................... | .................... | .................... |
| N | 1354 | hCoV-19/Wales/QEUH-9FB51F/2020\|EPI_ISL_600586\|2020 | .................... | .................... | .................... | .................... | .................... | .................... |
| N | 1355 | hCoV-19/Scotland/QEUH-9FA9AB/2020\|EPI_ISL_600587\|2 | .................... | .................... | .................... | .................... | .................... | .................... |
| N | 1356 | hCoV-19/Scotland/QEUH-9FB597/2020\|EPI_ISL_600588\|2 | .................... | .................... | .................... | .................... | .................... | .................... |
| N | 1357 | hCoV-19/Scotland/QEUH-9FB403/2020\|EPI_ISL_600589\|2 | .................... | .................... | .................... | .................... | .................... | .................... |
| N | 1358 | hCoV-19/Scotland/QEUH-9FB5B5/2020\|EPI_ISL_600595\|2 | .................... | .................... | .................... | .................... | .................... | .................... |
| N | 1359 | hCoV-19/Scotland/QEUH-9FAF9A/2020\|EPI_ISL_600596\|2 | .................... | .................... | .................... | .................... | .................... | .................... |
| N | 1360 | hCoV-19/Scotland/QEUH-9FB193/2020\|EPI_ISL_600597\|2 | .................... | .................... | .................... | .................... | .................... | .................... |
| N | 1361 | hCoV-19/Scotland/QEUH-9CFF25/2020\|EPI_ISL_601800\|2 | .................... | .................... | .................... | .................... | .................... | .................... |
| N | 1362 | hCoV-19/Scotland/QEUH-9CFF07/2020\|EPI_ISL_601801\|2 | .................... | .................... | .................... | .................... | .................... | .................... |
| N | 1363 | hCoV-19/Scotland/QEUH-9CFCE2/2020\|EPI_ISL_601804\|2 | .................... | .................... | .................... | .................... | .................... | .................... |
| N | 1364 | hCoV-19/Scotland/QEUH-9D0B4B/2020\|EPI_ISL_601806\|2 | .................... | .................... | .................... | .................... | .................... | .................... |
| N | 1365 | hCoV-19/Scotland/QEUH-9D0ECA/2020\|EPI_ISL_601807\|2 | .................... | .................... | .................... | .................... | .................... | .................... |
| N | 1366 | hCoV-19/Scotland/QEUH-9D0CC0/2020\|EPI_ISL_601809\|2 | .................... | .................... | .................... | .................... | .................... | .................... |
| N | 1367 | hCoV-19/Scotland/QEUH-9CFEBF/2020\|EPI_ISL_601810\|2 | .................... | .................... | .................... | .................... | .................... | .................... |
| N | 1368 | hCoV-19/Scotland/QEUH-9CFF43/2020\|EPI_ISL_601812\|2 | .................... | .................... | .................... | .................... | .................... | .................... |
| N | 1369 | hCoV-19/England/QEUH-9CFD0D/2020\|EPI_ISL_601813\|20 | .................... | .................... | .................... | .................... | .................... | .................... |
| N | 1370 | hCoV-19/Scotland/QEUH-9D0844/2020\|EPI_ISL_601814\|2 | .................... | .................... | .................... | .................... | ....T............... | .................... |
| N | 1371 | hCoV-19/Scotland/QEUH-9D03E9/2020\|EPI_ISL_601815\|2 | .................... | .................... | .................... | .................... | .................... | .................... |
| N | 1372 | hCoV-19/Scotland/QEUH-9D020A/2020\|EPI_ISL_601816\|2 | .................... | .................... | .................... | .................... | .................... | .................... |
| N | 1373 | hCoV-19/Scotland/QEUH-9D0792/2020\|EPI_ISL_601817\|2 | .................... | .................... | .................... | .................... | .................... | .................... |
| N | 1374 | hCoV-19/Scotland/QEUH-9CFE0A/2020\|EPI_ISL_601820\|2 | .................... | .................... | .................... | .................... | .................... | .................... |
| N | 1375 | hCoV-19/England/QEUH-9D00A6/2020\|EPI_ISL_601821\|20 | .................... | .................... | .................... | .................... | .................... | .................... |
| N | 1376 | hCoV-19/Scotland/QEUH-9D07A1/2020\|EPI_ISL_601822\|2 | .................... | .................... | .................... | .................... | .................... | .................... |
| N | 1377 | hCoV-19/Scotland/QEUH-9D045F/2020\|EPI_ISL_601823\|2 | .................... | .................... | .................... | .................... | .................... | .................... |
| N | 1378 | hCoV-19/Scotland/QEUH-9CFCF1/2020\|EPI_ISL_601824\|2 | .................... | .................... | .................... | .................... | ......T............. | .................... |
| N | 1379 | hCoV-19/Scotland/QEUH-9D11B1/2020\|EPI_ISL_601825\|2 | .................... | .................... | .................... | .................... | .................... | .................... |
| N | 1380 | hCoV-19/Scotland/QEUH-9CFC4C/2020\|EPI_ISL_601826\|2 | .................... | .................... | .................... | .................... | .................... | .................... |
| N | 1381 | hCoV-19/Scotland/QEUH-9D0C2A/2020\|EPI_ISL_601827\|2 | .................... | .................... | .................... | .................... | .................... | .................... |
| N | 1382 | hCoV-19/Scotland/QEUH-9D0228/2020\|EPI_ISL_601828\|2 | .................... | .................... | .................... | .................... | .................... | .................... |
| N | 1383 | hCoV-19/Scotland/QEUH-9D0B1E/2020\|EPI_ISL_601829\|2 | .................... | .................... | .................... | .................... | .................... | .................... |
| N | 1384 | hCoV-19/Scotland/QEUH-9CFEFB/2020\|EPI_ISL_601830\|2 | .................... | .................... | .................... | .................... | .................... | .................... |
| N | 1385 | hCoV-19/Scotland/QEUH-9D03CB/2020\|EPI_ISL_601831\|2 | .................... | .................... | .................... | .................... | .................... | .................... |
| N | 1386 | hCoV-19/Scotland/QEUH-9D0219/2020\|EPI_ISL_601832\|2 | .................... | .................... | .................... | .................... | .................... | .................... |
| N | 1387 | hCoV-19/Scotland/QEUH-9CFCD3/2020\|EPI_ISL_601833\|2 | .................... | .................... | .................... | .................... | .................... | .................... |
| N | 1388 | hCoV-19/Scotland/QEUH-9D0808/2020\|EPI_ISL_601835\|2 | .................... | .................... | .................... | .................... | .................... | .................... |
| N | 1389 | hCoV-19/Scotland/QEUH-9D0C0C/2020\|EPI_ISL_601837\|2 | .................... | .................... | .................... | .................... | .................... | .................... |
| N | 1390 | hCoV-19/Scotland/QEUH-9CFDFE/2020\|EPI_ISL_601838\|2 | .................... | .................... | .................... | .................... | .................... | .................... |
| N | 1391 | hCoV-19/Scotland/QEUH-9D08F9/2020\|EPI_ISL_601839\|2 | .................... | .................... | .................... | .................... | .................... | .................... |
| N | 1392 | hCoV-19/Scotland/QEUH-9D0CA2/2020\|EPI_ISL_601840\|2 | .................... | .................... | .................... | .................... | .................... | .................... |
| N | 1393 | hCoV-19/Scotland/QEUH-9D0404/2020\|EPI_ISL_601842\|2 | .................... | .................... | .................... | .................... | .................... | .................... |
| N | 1394 | hCoV-19/Scotland/QEUH-9D0853/2020\|EPI_ISL_601843\|2 | .................... | .................... | .................... | .................... | .................... | .................... |
| N | 1395 | hCoV-19/Scotland/QEUH-9CFEDD/2020\|EPI_ISL_601844\|2 | .................... | .................... | .................... | .................... | .................... | .................... |
| N | 1396 | hCoV-19/Scotland/QEUH-9CE278/2020\|EPI_ISL_601889\|2 | .................... | .................... | .................... | .................... | .................... | .................... |
| N | 1397 | hCoV-19/Scotland/QEUH-9CE9CE/2020\|EPI_ISL_601890\|2 | .................... | .................... | .................... | .................... | .................... | .................... |
| N | 1398 | hCoV-19/Scotland/QEUH-9CE24B/2020\|EPI_ISL_601891\|2 | .................... | .................... | .................... | .................... | .................... | .................... |
| N | 1399 | hCoV-19/Scotland/QEUH-9CE30C/2020\|EPI_ISL_601892\|2 | .................... | .................... | .................... | .................... | .................... | .................... |
| N | 1400 | hCoV-19/Scotland/QEUH-9CEFEA/2020\|EPI_ISL_601893\|2 | .................... | .................... | .................... | .................... | .................... | .................... |
| N | 1401 | hCoV-19/Scotland/QEUH-9CE2B4/2020\|EPI_ISL_601894\|2 | .................... | .................... | .................... | .................... | .................... | .................... |
| N | 1402 | hCoV-19/Scotland/QEUH-9CF3B0/2020\|EPI_ISL_601895\|2 | .................... | .................... | .................... | .................... | .................... | .................... |
| N | 1403 | hCoV-19/Scotland/QEUH-9CF514/2020\|EPI_ISL_601896\|2 | .................... | .................... | .................... | .................... | .................... | .................... |
| N | 1404 | hCoV-19/Scotland/QEUH-9CE542/2020\|EPI_ISL_601897\|2 | .................... | .................... | .................... | .................... | .................... | .................... |
| N | 1405 | hCoV-19/Scotland/QEUH-9CEA34/2020\|EPI_ISL_601898\|2 | .................... | .................... | .................... | .................... | .................... | .................... |
| N | 1406 | hCoV-19/Scotland/QEUH-9CE296/2020\|EPI_ISL_601899\|2 | .................... | .................... | .................... | .................... | .................... | .................... |
| N | 1407 | hCoV-19/USA/MI-QDX-1712/2020\|EPI_ISL_604040\|2020-0 | .................... | .................... | .................... | .................... | .................... | .................... |
| N | 1408 | hCoV-19/USA/MI-QDX-1723/2020\|EPI_ISL_604041\|2020-0 | .................... | .................... | .................... | .................... | .................... | .................... |
| N | 1409 | hCoV-19/USA/MI-QDX-1726/2020\|EPI_ISL_604042\|2020-0 | .................... | .................... | .................... | .................... | .................... | .................... |
| N | 1410 | hCoV-19/USA/MI-QDX-1728/2020\|EPI_ISL_604043\|2020-0 | .................... | .................... | .................... | .................... | .................... | .................... |
| N | 1411 | hCoV-19/USA/MI-QDX-1736/2020\|EPI_ISL_604044\|2020-0 | .................... | .................... | .................... | .................... | .................... | .................... |
| N | 1412 | hCoV-19/USA/MI-QDX-1738/2020\|EPI_ISL_604045\|2020-0 | .................... | .................... | .................... | .................... | .................... | .................... |

| | | | | | | | |
|---|---|---|---|---|---|---|---|
| N | 1413 hCoV-19/USA/MI-QDX-1743/2020\|EPI_ISL_604046\|2020-0 | ···················· | ···················· | ···················· | ···················· | ···················· | ···················· |
| N | 1414 hCoV-19/USA/MI-QDX-1746/2020\|EPI_ISL_604047\|2020-0 | ···················· | ···················· | ···················· | ···················· | ···················· | ···················· |
| N | 1415 hCoV-19/USA/MI-QDX-1747/2020\|EPI_ISL_604048\|2020-0 | ···················· | ···················· | ···················· | ···················· | ···················· | ···················· |
| N | 1416 hCoV-19/USA/MI-QDX-1748/2020\|EPI_ISL_604049\|2020-0 | ···················· | ···················· | ···················· | ···················· | ···················· | ···················· |
| N | 1417 hCoV-19/USA/MI-QDX-1751/2020\|EPI_ISL_604050\|2020-0 | ···················· | ···················· | ···················· | ···················· | ···················· | ···················· |
| N | 1418 hCoV-19/USA/MI-QDX-1752/2020\|EPI_ISL_604051\|2020-0 | ···················· | ···················· | ···················· | ···················· | ···················· | ···················· |
| N | 1419 hCoV-19/USA/MI-QDX-1755/2020\|EPI_ISL_604052\|2020-0 | ···················· | ···················· | ···················· | ···················· | ···················· | ···················· |
| N | 1420 hCoV-19/USA/MI-QDX-1757/2020\|EPI_ISL_604053\|2020-0 | ···················· | ···················· | ···················· | ···················· | ···················· | ···················· |
| N | 1421 hCoV-19/USA/MI-QDX-1761/2020\|EPI_ISL_604054\|2020-0 | ···················· | ···················· | ···················· | ···················· | ···················· | ···················· |
| N | 1422 hCoV-19/USA/MI-QDX-1767/2020\|EPI_ISL_604055\|2020-0 | ···················· | ···················· | ···················· | ···················· | ···················· | ···················· |
| N | 1423 hCoV-19/USA/MI-QDX-1769/2020\|EPI_ISL_604056\|2020-0 | ···················· | ···················· | ···················· | ···················· | ···················· | ···················· |
| N | 1424 hCoV-19/USA/MI-QDX-1770/2020\|EPI_ISL_604057\|2020-0 | ···················· | ·········A·········· | ···················· | ···················· | ···················· | ···················· |
| N | 1425 hCoV-19/USA/MI-QDX-1772/2020\|EPI_ISL_604058\|2020-0 | ···················· | ···················· | ···················· | ···················· | ···················· | ···················· |
| N | 1426 hCoV-19/USA/MI-QDX-1780/2020\|EPI_ISL_604059\|2020-0 | ···················· | ···················· | ···················· | ···················· | ···················· | ···················· |
| N | 1427 hCoV-19/USA/MN-QDX-1701/2020\|EPI_ISL_604060\|2020-0 | ···················· | ···················· | ···················· | ···················· | ···················· | ···················· |
| N | 1428 hCoV-19/USA/MN-QDX-1702/2020\|EPI_ISL_604061\|2020-0 | ···················· | ···················· | ···················· | ···················· | ···················· | ···················· |
| N | 1429 hCoV-19/USA/MN-QDX-1706/2020\|EPI_ISL_604062\|2020-0 | ···················· | ···················· | ···················· | ···················· | ···················· | ···················· |
| N | 1430 hCoV-19/USA/MN-QDX-1708/2020\|EPI_ISL_604063\|2020-0 | ···················· | ···················· | ···················· | ···················· | ···················· | ···················· |
| N | 1431 hCoV-19/USA/MN-QDX-1710/2020\|EPI_ISL_604064\|2020-0 | ···················· | ···················· | ···················· | ···················· | ···················· | ···················· |
| N | 1432 hCoV-19/USA/MN-QDX-1711/2020\|EPI_ISL_604065\|2020-0 | ···················· | ···················· | ···················· | ···················· | ···················· | ···················· |
| N | 1433 hCoV-19/USA/MN-QDX-1714/2020\|EPI_ISL_604066\|2020-0 | ···················· | ···················· | ···················· | ···················· | ···················· | ···················· |
| N | 1434 hCoV-19/USA/MN-QDX-1715/2020\|EPI_ISL_604067\|2020-0 | ···················· | ···················· | ···················· | ···················· | ···················· | ···················· |
| N | 1435 hCoV-19/USA/MN-QDX-1716/2020\|EPI_ISL_604068\|2020-0 | ···················· | ···················· | ···················· | ···················· | ···················· | ···················· |
| N | 1436 hCoV-19/USA/MN-QDX-1718/2020\|EPI_ISL_604069\|2020-0 | ···················· | ···················· | ···················· | ···················· | ···················· | ···················· |
| N | 1437 hCoV-19/USA/MN-QDX-1721/2020\|EPI_ISL_604070\|2020-0 | ···················· | ···················· | ···················· | ···················· | ···················· | ···················· |
| N | 1438 hCoV-19/USA/MN-QDX-1722/2020\|EPI_ISL_604071\|2020-0 | ···················· | ···················· | ···················· | ···················· | ···················· | ···················· |
| N | 1439 hCoV-19/USA/MN-QDX-1724/2020\|EPI_ISL_604072\|2020-0 | ···················· | ···················· | ···················· | ···················· | ···················· | ···················· |
| N | 1440 hCoV-19/USA/MN-QDX-1730/2020\|EPI_ISL_604073\|2020-0 | ···················· | ···················· | ···················· | ···················· | ···················· | ···················· |
| N | 1441 hCoV-19/USA/MN-QDX-1732/2020\|EPI_ISL_604074\|2020-0 | ···················· | ···················· | ···················· | ···················· | ···················· | ···················· |
| N | 1442 hCoV-19/USA/MN-QDX-1733/2020\|EPI_ISL_604075\|2020-0 | ···················· | ······A············· | ···················· | ···················· | ···················· | ···················· |
| N | 1443 hCoV-19/USA/MN-QDX-1734/2020\|EPI_ISL_604076\|2020-0 | ···················· | ···················· | ···················· | ···················· | ···················· | ···················· |
| N | 1444 hCoV-19/USA/MN-QDX-1737/2020\|EPI_ISL_604077\|2020-0 | ···················· | ···················· | ···················· | ···················· | ···················· | ···················· |
| N | 1445 hCoV-19/USA/MN-QDX-1739/2020\|EPI_ISL_604078\|2020-0 | ···················· | ···················· | ···················· | ···················· | ···················· | ···················· |
| N | 1446 hCoV-19/USA/MN-QDX-1740/2020\|EPI_ISL_604079\|2020-0 | ···················· | ···················· | ···················· | ···················· | ···················· | ···················· |
| N | 1447 hCoV-19/USA/MN-QDX-1741/2020\|EPI_ISL_604080\|2020-0 | ···················· | ···················· | ···················· | ···················· | ···················· | ···················· |
| N | 1448 hCoV-19/USA/MN-QDX-1744/2020\|EPI_ISL_604081\|2020-0 | ···················· | ···················· | ···················· | ···················· | ···················· | ···················· |
| N | 1449 hCoV-19/USA/MN-QDX-1750/2020\|EPI_ISL_604082\|2020-0 | ···················· | ···················· | ···················· | ···················· | ···················· | ···················· |
| N | 1450 hCoV-19/USA/MN-QDX-1753/2020\|EPI_ISL_604083\|2020-0 | ···················· | ···················· | ···················· | ···················· | ···················· | ···················· |
| N | 1451 hCoV-19/USA/MN-QDX-1754/2020\|EPI_ISL_604084\|2020-0 | ···················· | ···················· | ···················· | ···················· | ···················· | ···················· |
| N | 1452 hCoV-19/USA/MN-QDX-1760/2020\|EPI_ISL_604085\|2020-0 | ···················· | ···················· | ···················· | ···················· | ···················· | ···················· |
| N | 1453 hCoV-19/USA/MN-QDX-1763/2020\|EPI_ISL_604086\|2020-0 | ···················· | ···················· | ···················· | ···················· | ···················· | ···················· |
| N | 1454 hCoV-19/USA/MN-QDX-1766/2020\|EPI_ISL_604087\|2020-0 | ···················· | ···················· | ···················· | ···················· | ···················· | ···················· |
| N | 1455 hCoV-19/USA/MN-QDX-1768/2020\|EPI_ISL_604088\|2020-0 | ···················· | ···················· | ···················· | ···················· | ···················· | ···················· |
| N | 1456 hCoV-19/USA/MN-QDX-1774/2020\|EPI_ISL_604089\|2020-0 | ···················· | ···················· | ···················· | ···················· | ···················· | ···················· |
| N | 1457 hCoV-19/USA/MN-QDX-1778/2020\|EPI_ISL_604090\|2020-0 | ···················· | ···················· | ···················· | ···················· | ···················· | ···················· |
| N | 1458 hCoV-19/USA/MN-QDX-1779/2020\|EPI_ISL_604091\|2020-0 | ···················· | ···················· | ···················· | ···················· | ···················· | ···················· |
| N | 1459 hCoV-19/USA/OH-QDX-1759/2020\|EPI_ISL_604092\|2020-0 | ···················· | ···················· | ···················· | ···················· | ···················· | ···················· |
| N | 1460 hCoV-19/USA/TX-QDX-1764/2020\|EPI_ISL_604093\|2020-0 | ···················· | ···················· | ···················· | ···················· | ···················· | ···················· |
| N | 1461 hCoV-19/USA/AZ-QDX-1798/2020\|EPI_ISL_604094\|2020-0 | ···················· | ···················· | ···················· | ···················· | ···················· | ···················· |
| N | 1462 hCoV-19/USA/AZ-QDX-1802/2020\|EPI_ISL_604095\|2020-0 | ···················· | ···················· | ···················· | ···················· | ···················· | ···················· |
| N | 1463 hCoV-19/USA/AZ-QDX-1819/2020\|EPI_ISL_604096\|2020-0 | ···················· | ···················· | ···················· | ···················· | ···················· | ···················· |
| N | 1464 hCoV-19/USA/AZ-QDX-1840/2020\|EPI_ISL_604097\|2020-0 | ···················· | ···················· | ···················· | ···················· | ···················· | ···················· |
| N | 1465 hCoV-19/USA/CA-QDX-1784/2020\|EPI_ISL_604098\|2020-0 | ···················· | ···················· | ···················· | ···················· | ···················· | ···················· |
| N | 1466 hCoV-19/USA/CA-QDX-1792/2020\|EPI_ISL_604099\|2020-0 | ···················· | ···················· | ···················· | ···················· | ···················· | ···················· |
| N | 1467 hCoV-19/USA/CA-QDX-1810/2020\|EPI_ISL_604100\|2020-0 | ···················· | ···················· | ···················· | ···················· | ···················· | ···················· |
| N | 1468 hCoV-19/USA/CA-QDX-1813/2020\|EPI_ISL_604101\|2020-0 | ···················· | ···················· | ···················· | ···················· | ···················· | ···················· |
| N | 1469 hCoV-19/USA/CA-QDX-1821/2020\|EPI_ISL_604102\|2020-0 | ···················· | ···················· | ···················· | ···················· | ···················· | ···················· |
| N | 1470 hCoV-19/USA/CA-QDX-1824/2020\|EPI_ISL_604103\|2020-0 | ···················· | ···················· | ···················· | ···················· | ···················· | ···················· |
| N | 1471 hCoV-19/USA/CA-QDX-1855/2020\|EPI_ISL_604104\|2020-0 | ···················· | ···················· | ···················· | ···················· | ···················· | ···················· |
| N | 1472 hCoV-19/USA/CT-QDX-1837/2020\|EPI_ISL_604105\|2020-0 | ···················· | ···················· | ···················· | ···················· | ···················· | ···················· |
| N | 1473 hCoV-19/USA/FL-QDX-1820/2020\|EPI_ISL_604106\|2020-0 | ···················· | ···················· | ···················· | ···················· | ···················· | ···················· |
| N | 1474 hCoV-19/USA/MA-QDX-1782/2020\|EPI_ISL_604107\|2020-0 | ···················· | ···················· | ···················· | ···················· | ···················· | ···················· |
| N | 1475 hCoV-19/USA/MA-QDX-1791/2020\|EPI_ISL_604108\|2020-0 | ···················· | ···················· | ···················· | ···················· | ···················· | ···················· |
| N | 1476 hCoV-19/USA/MA-QDX-1817/2020\|EPI_ISL_604109\|2020-0 | ···················· | ···················· | ···················· | ···················· | ···················· | ···················· |
| N | 1477 hCoV-19/USA/MA-QDX-1826/2020\|EPI_ISL_604110\|2020-0 | ···················· | ···················· | ···················· | ···················· | ···················· | ···················· |
| N | 1478 hCoV-19/USA/MA-QDX-1862/2020\|EPI_ISL_604111\|2020-0 | ···················· | ···················· | ···················· | ···················· | ···················· | ···················· |
| N | 1479 hCoV-19/USA/ME-QDX-1803/2020\|EPI_ISL_604112\|2020-0 | ···················· | ···················· | ···················· | ···················· | ···················· | ···················· |
| N | 1480 hCoV-19/USA/ME-QDX-1814/2020\|EPI_ISL_604113\|2020-0 | ···················· | ···················· | ···················· | ···················· | ···················· | ···················· |
| N | 1481 hCoV-19/USA/ME-QDX-1857/2020\|EPI_ISL_604114\|2020-0 | ···················· | ···················· | ···················· | ···················· | ···················· | ···················· |
| N | 1482 hCoV-19/USA/UT-QDX-1781/2020\|EPI_ISL_604115\|2020-0 | ···················· | ···················· | ···················· | ···················· | ···················· | ···················· |
| N | 1483 hCoV-19/USA/UT-QDX-1783/2020\|EPI_ISL_604116\|2020-0 | ···················· | ···················· | ···················· | ···················· | ···················· | ···················· |
| N | 1484 hCoV-19/USA/UT-QDX-1785/2020\|EPI_ISL_604117\|2020-0 | ···················· | ···················· | ···················· | ···················· | ···················· | ···················· |
| N | 1485 hCoV-19/USA/UT-QDX-1786/2020\|EPI_ISL_604118\|2020-0 | ···················· | ···················· | ···················· | ···················· | ···················· | ···················· |
| N | 1486 hCoV-19/USA/UT-QDX-1787/2020\|EPI_ISL_604119\|2020-0 | ···················· | ···················· | ···················· | ···················· | ···················· | ···················· |
| N | 1487 hCoV-19/USA/UT-UPHL-2009698/2020\|EPI_ISL_605372\|20 | ···················· | ···················· | ···················· | ···················· | ···················· | ···················· |
| N | 1488 hCoV-19/USA/UT-UPHL-2009699/2020\|EPI_ISL_605373\|20 | ···················· | ···················· | ···················· | ···················· | ···················· | ···················· |
| N | 1489 hCoV-19/USA/UT-UPHL-2009700/2020\|EPI_ISL_605374\|20 | ···················· | ···················· | ···················· | ···················· | ···················· | ···················· |
| N | 1490 hCoV-19/USA/UT-UPHL-2009702/2020\|EPI_ISL_605375\|20 | ···················· | ·A·················· | ···················· | ···················· | ···················· | ···················· |
| N | 1491 hCoV-19/USA/UT-UPHL-2009703/2020\|EPI_ISL_605376\|20 | ···················· | ···················· | ···················· | ···················· | ···················· | ···················· |
| N | 1492 hCoV-19/USA/UT-UPHL-2009704/2020\|EPI_ISL_605377\|20 | ···················· | ···················· | ···················· | ···················· | ···················· | ···················· |

| | | | | | | | | |
|---|---|---|---|---|---|---|---|---|
| N | 1493 | hCoV-19/USA/UT-UPHL-2009705/2020\|EPI_ISL_605378\|20 | ···················· | ··G················· | ···················· | ···················· | ···················· | ···················· |
| N | 1494 | hCoV-19/USA/UT-UPHL-2009706/2020\|EPI_ISL_605379\|20 | ···················· | ···················· | ···················· | ···················· | ···················· | ···················· |
| N | 1495 | hCoV-19/USA/UT-UPHL-2009709/2020\|EPI_ISL_605382\|20 | ···················· | ···················· | ···················· | ···················· | ···················· | ···················· |
| N | 1496 | hCoV-19/USA/UT-UPHL-2009710/2020\|EPI_ISL_605383\|20 | ···················· | ···················· | ···················· | ···················· | ···················· | ···················· |
| N | 1497 | hCoV-19/USA/UT-UPHL-2009711/2020\|EPI_ISL_605384\|20 | ···················· | ···················· | ···················· | ···················· | ···················· | ···················· |
| N | 1498 | hCoV-19/USA/UT-UPHL-2009712/2020\|EPI_ISL_605385\|20 | ···················· | ···················· | ···················· | ···················· | ···················· | ···················· |
| N | 1499 | hCoV-19/USA/UT-UPHL-2009713/2020\|EPI_ISL_605386\|20 | ···················· | ···················· | ···················· | ···················· | ···················· | ···················· |
| N | 1500 | hCoV-19/USA/UT-UPHL-2009716/2020\|EPI_ISL_605388\|20 | ···················· | ···················· | ···················· | ···················· | ···················· | ···················· |
| N | 1501 | hCoV-19/USA/UT-UPHL-2009717/2020\|EPI_ISL_605389\|20 | ···················· | ···················· | ···················· | ···················· | ···················· | ···················· |
| N | 1502 | hCoV-19/USA/UT-UPHL-2009718/2020\|EPI_ISL_605390\|20 | ···················· | ···················· | ···················· | ···················· | ···················· | ···················· |
| N | 1503 | hCoV-19/USA/UT-UPHL-2009719/2020\|EPI_ISL_605391\|20 | ···················· | ···················· | ···················· | ···················· | ···················· | ···················· |
| N | 1504 | hCoV-19/USA/UT-UPHL-2009720/2020\|EPI_ISL_605392\|20 | ···················· | ···················· | ···················· | ···················· | ···················· | ···················· |
| N | 1505 | hCoV-19/USA/UT-UPHL-2009721/2020\|EPI_ISL_605393\|20 | ···················· | ···················· | ···················· | ···················· | ···················· | ···················· |
| N | 1506 | hCoV-19/USA/UT-UPHL-2009724/2020\|EPI_ISL_605396\|20 | ···················· | ·A·················· | ···················· | ···················· | ···················· | ···················· |
| N | 1507 | hCoV-19/USA/UT-UPHL-2009726/2020\|EPI_ISL_605398\|20 | ···················· | ···················· | ···················· | ···················· | ···················· | ···················· |
| N | 1508 | hCoV-19/USA/UT-UPHL-2009727/2020\|EPI_ISL_605399\|20 | ···················· | ···················· | ···················· | ···················· | ···················· | ···················· |
| N | 1509 | hCoV-19/USA/WI-UW-1542/2020\|EPI_ISL_605420\|2020-09 | ···················· | ···················· | ···················· | ···················· | ···················· | ···················· |
| N | 1510 | hCoV-19/USA/WI-UW-1543/2020\|EPI_ISL_605421\|2020-09 | ···················· | ···················· | ···················· | ···················· | ···················· | ···················· |
| N | 1511 | hCoV-19/USA/WI-UW-1544/2020\|EPI_ISL_605422\|2020-09 | ···················· | ···················· | ···················· | ···················· | ···················· | ···················· |
| N | 1512 | hCoV-19/USA/WI-UW-1545/2020\|EPI_ISL_605423\|2020-09 | ···················· | ···················· | ···················· | ···················· | ···················· | ···················· |
| N | 1513 | hCoV-19/USA/WI-UW-1546/2020\|EPI_ISL_605424\|2020-09 | ···················· | ···················· | ···················· | ···················· | ···················· | ···················· |
| N | 1514 | hCoV-19/USA/WI-UW-1547/2020\|EPI_ISL_605425\|2020-09 | ···················· | ···················· | ···················· | ···················· | ···················· | ···················· |
| N | 1515 | hCoV-19/USA/WI-UW-1549/2020\|EPI_ISL_605427\|2020-09 | ···················· | ···················· | ···················· | ···················· | ···················· | ···················· |
| N | 1516 | hCoV-19/USA/WI-UW-1550/2020\|EPI_ISL_605428\|2020-09 | ···················· | ···················· | ···················· | ···················· | ···················· | ···················· |
| N | 1517 | hCoV-19/USA/WI-UW-1551/2020\|EPI_ISL_605429\|2020-09 | ···················· | ···················· | ···················· | ···················· | ···················· | ···················· |
| N | 1521 | hCoV-19/USA/WI-UW-1555/2020\|EPI_ISL_605433\|2020-09 | ···················· | ···················· | ···················· | ···················· | ···················· | ···················· |
| N | 1527 | hCoV-19/USA/WI-UW-1562/2020\|EPI_ISL_605440\|2020-09 | ···················· | ···················· | ···················· | ···················· | ···················· | ···················· |
| N | 1529 | hCoV-19/USA/WI-UW-1564/2020\|EPI_ISL_605442\|2020-09 | ···················· | ···················· | ···················· | ···················· | ···················· | ···················· |
| N | 1532 | hCoV-19/USA/WI-UW-1567/2020\|EPI_ISL_605445\|2020-09 | ···················· | ···················· | ···················· | ···················· | ···················· | ···················· |
| N | 1535 | hCoV-19/USA/WI-UW-1570/2020\|EPI_ISL_605448\|2020-09 | ···················· | ···················· | ···················· | ···················· | ···················· | ···················· |
| N | 1537 | hCoV-19/Australia/WA137/2020\|EPI_ISL_596700\|2020-0 | ···················· | ···················· | ···················· | ···················· | ···················· | ···················· |
| N | 1538 | hCoV-19/Australia/WA138/2020\|EPI_ISL_596701\|2020-0 | ···················· | ···················· | ···················· | ···················· | ···················· | ···················· |
| N | 1539 | hCoV-19/Australia/WA139/2020\|EPI_ISL_596702\|2020-0 | ···················· | ···················· | ···················· | ···················· | ···················· | ···················· |
| N | 1540 | hCoV-19/Australia/WA140/2020\|EPI_ISL_596703\|2020-0 | ···················· | ···················· | ···················· | ···················· | ···················· | ···················· |
| N | 1541 | hCoV-19/Australia/WA141/2020\|EPI_ISL_596704\|2020-0 | ···················· | ···················· | ···················· | ···················· | ···················· | ···················· |
| N | 1542 | hCoV-19/Australia/WA142/2020\|EPI_ISL_596705\|2020-0 | ···················· | ···················· | ···················· | ···················· | ···················· | ···················· |
| N | 1543 | hCoV-19/Australia/WA143/2020\|EPI_ISL_596706\|2020-0 | ···················· | ···················· | ···················· | ···················· | ···················· | ···················· |
| N | 1544 | hCoV-19/Australia/WA144/2020\|EPI_ISL_596707\|2020-0 | ···················· | ···················· | ···················· | ···················· | ···················· | ···················· |
| N | 1545 | hCoV-19/Australia/WA145/2020\|EPI_ISL_596708\|2020-0 | ···················· | ···················· | ···················· | ···················· | ···················· | ···················· |
| N | 1546 | hCoV-19/Australia/WA146/2020\|EPI_ISL_596709\|2020-0 | ···················· | ···················· | ···················· | ···················· | ···················· | ···················· |
| N | 1547 | hCoV-19/Australia/WA147/2020\|EPI_ISL_596710\|2020-0 | ···················· | ···················· | ···················· | ···················· | ···················· | ···················· |
| N | 1548 | hCoV-19/Australia/WA148/2020\|EPI_ISL_596711\|2020-0 | ···················· | ···················· | ···················· | ···················· | ···················· | ···················· |
| N | 1550 | hCoV-19/Australia/WA150/2020\|EPI_ISL_596713\|2020-0 | ···················· | ···················· | ···················· | ···················· | ···················· | ···················· |
| N | 1551 | hCoV-19/England/QEUH-9FFC34/2020\|EPI_ISL_600447\|20 | ···················· | ···················· | ···················· | ···················· | ···················· | ···················· |
| N | 1552 | hCoV-19/Scotland/QEUH-9FFE2F/2020\|EPI_ISL_600481\|2 | ···················· | ···················· | ···················· | ···················· | ···················· | ···················· |
| N | 1553 | hCoV-19/England/QEUH-9FF311/2020\|EPI_ISL_600482\|20 | ···················· | ···················· | ···················· | ···················· | ···················· | ···················· |
| N | 1554 | hCoV-19/England/QEUH-A00060/2020\|EPI_ISL_600483\|20 | ···················· | ·A·················· | ···················· | ···················· | ···················· | ···················· |
| N | 1555 | hCoV-19/England/QEUH-9FFAA3/2020\|EPI_ISL_600485\|20 | ···················· | ···················· | ···················· | ···················· | ···················· | ···················· |
| N | 1556 | hCoV-19/England/QEUH-9FFB91/2020\|EPI_ISL_600486\|20 | ···················· | ···················· | ···················· | ···················· | ···················· | ···················· |
| N | 1557 | hCoV-19/Scotland/QEUH-9FF663/2020\|EPI_ISL_600488\|2 | ···················· | ···················· | ···················· | ···················· | ···················· | ···················· |
| N | 1558 | hCoV-19/England/QEUH-9FF126/2020\|EPI_ISL_600490\|20 | ···················· | ···················· | ···················· | ···················· | ···················· | ···················· |
| N | 1559 | hCoV-19/England/QEUH-9FF627/2020\|EPI_ISL_600491\|20 | ···················· | ·A·················· | ···················· | ···················· | ···················· | ···················· |
| N | 1560 | hCoV-19/England/QEUH-9FFFE0/2020\|EPI_ISL_600493\|20 | ···················· | ···················· | ···················· | ···················· | ···················· | ···················· |
| N | 1561 | hCoV-19/Scotland/QEUH-A007B6/2020\|EPI_ISL_600494\|2 | ···················· | ···················· | ···················· | ···················· | ···················· | ···················· |
| N | 1562 | hCoV-19/England/QEUH-A006F5/2020\|EPI_ISL_600496\|20 | ···················· | ···················· | ···················· | ···················· | ···················· | ···················· |
| N | 1563 | hCoV-19/England/QEUH-A00428/2020\|EPI_ISL_600497\|20 | ···················· | ···················· | ···················· | ···················· | ···················· | ···················· |
| N | 1564 | hCoV-19/England/QEUH-A00367/2020\|EPI_ISL_600499\|20 | ···················· | ·A·················· | ···················· | ···················· | ···················· | ···················· |
| N | 1565 | hCoV-19/England/QEUH-9FF223/2020\|EPI_ISL_600514\|20 | ···················· | ···················· | ···················· | ···················· | ···················· | ···················· |
| N | 1566 | hCoV-19/Scotland/QEUH-9FFCE9/2020\|EPI_ISL_600516\|2 | ···················· | ···················· | ···················· | ···················· | ···················· | ···················· |
| N | 1567 | hCoV-19/England/QEUH-A00570/2020\|EPI_ISL_600518\|20 | ···················· | ···················· | ···················· | ···················· | ···················· | ···················· |
| N | 1568 | hCoV-19/England/QEUH-9FF8E5/2020\|EPI_ISL_600520\|20 | ···················· | ···················· | ···················· | ···················· | ···················· | ···················· |
| N | 1569 | hCoV-19/Wales/ALDP-9FDBDF/2020\|EPI_ISL_600521\|2020 | ···················· | ···················· | ···················· | ···················· | ···················· | ···················· |
| N | 1570 | hCoV-19/Scotland/ALDP-9FDD42/2020\|EPI_ISL_600523\|2 | ···················· | ···················· | ···················· | ···················· | ···················· | ···················· |
| N | 1571 | hCoV-19/Wales/ALDP-9FDB66/2020\|EPI_ISL_600525\|2020 | ···················· | ···················· | ···················· | ···················· | ···················· | ···················· |
| N | 1572 | hCoV-19/Wales/ALDP-9FDCCD/2020\|EPI_ISL_600526\|2020 | ···················· | ···················· | ···················· | ···················· | ···················· | ···················· |
| N | 1573 | hCoV-19/Wales/ALDP-9FDB93/2020\|EPI_ISL_600528\|2020 | ···················· | ···················· | ···················· | ···················· | ···················· | ···················· |
| N | 1574 | hCoV-19/Wales/ALDP-9FDBA2/2020\|EPI_ISL_600529\|2020 | ···················· | ···················· | ···················· | ···················· | ···················· | ···················· |
| N | 1575 | hCoV-19/Wales/ALDP-9FDC72/2020\|EPI_ISL_600531\|2020 | ···················· | ···················· | ···················· | ···················· | ···················· | ···················· |
| N | 1576 | hCoV-19/Wales/ALDP-9FDCAF/2020\|EPI_ISL_600532\|2020 | ···················· | ···················· | ···················· | ···················· | ···················· | ···················· |
| N | 1577 | hCoV-19/Wales/ALDP-9FDC36/2020\|EPI_ISL_600534\|2020 | ···················· | ···················· | ···················· | ···················· | ···················· | ···················· |
| N | 1578 | hCoV-19/Scotland/ALDP-9FDDAC/2020\|EPI_ISL_600536\|2 | ···················· | ···················· | ···················· | ···················· | ···················· | ···················· |
| N | 1579 | hCoV-19/Wales/ALDP-9FC781/2020\|EPI_ISL_600537\|2020 | ···················· | ···················· | ···················· | ···················· | ···················· | ···················· |
| N | 1580 | hCoV-19/Scotland/QEUH-9FA98D/2020\|EPI_ISL_600539\|2 | ···················· | ···················· | ···················· | ···················· | ···················· | ···················· |
| N | 1581 | hCoV-19/Scotland/QEUH-9FB5E2/2020\|EPI_ISL_600540\|2 | ···················· | ···················· | ···················· | ···················· | ···················· | ···················· |
| N | 1582 | hCoV-19/Scotland/QEUH-9FA5B6/2020\|EPI_ISL_600542\|2 | ···················· | ···················· | ···················· | ···················· | ···················· | ···················· |
| N | 1583 | hCoV-19/Scotland/QEUH-9FA9F6/2020\|EPI_ISL_600544\|2 | ···················· | ···················· | ···················· | ···················· | ···················· | ···················· |
| N | 1584 | hCoV-19/Scotland/QEUH-9FA8EA/2020\|EPI_ISL_600545\|2 | ···················· | ···················· | ···················· | ···················· | ···················· | ···················· |
| N | 1585 | hCoV-19/Scotland/QEUH-9FB3BB/2020\|EPI_ISL_600547\|2 | ···················· | ···················· | ···················· | ···················· | ···················· | ···················· |
| N | 1586 | hCoV-19/Scotland/QEUH-9FB45E/2020\|EPI_ISL_600548\|2 | ···················· | ···················· | ···················· | ···················· | ···················· | ···················· |
| N | 1587 | hCoV-19/Scotland/QEUH-9FAD90/2020\|EPI_ISL_600550\|2 | ···················· | ···················· | ···················· | ···················· | ···················· | ···················· |

| | | | | | | | | |
|---|---|---|---|---|---|---|---|---|
| N | 1588 | hCoV-19/Scotland/QEUH-9FB37F/2020\|EPI_ISL_600551\|2 | ···················· | ···················· | ···················· | ···················· | ···················· | ···················· |
| N | 1589 | hCoV-19/Scotland/QEUH-9FB351/2020\|EPI_ISL_600553\|2 | ···················· | ···················· | ···················· | ···················· | ···················· | ···················· |
| N | 1590 | hCoV-19/Scotland/QEUH-9FA862/2020\|EPI_ISL_600555\|2 | ···················· | ···················· | ···················· | ···················· | ···················· | ···················· |
| N | 1591 | hCoV-19/Scotland/QEUH-9FB421/2020\|EPI_ISL_600556\|2 | ···················· | ···················· | ···················· | ···················· | ···················· | ···················· |
| N | 1592 | hCoV-19/Scotland/QEUH-9FB5D3/2020\|EPI_ISL_600590\|2 | ···················· | ···················· | ···················· | ···················· | ···················· | ···················· |
| N | 1593 | hCoV-19/Scotland/QEUH-9FB3AC/2020\|EPI_ISL_600591\|2 | ···················· | ···················· | ···················· | ···················· | ···················· | ···················· |
| N | 1594 | hCoV-19/Scotland/QEUH-9FA9BA/2020\|EPI_ISL_600592\|2 | ···················· | ···················· | ···················· | ···················· | ···················· | ···················· |
| N | 1595 | hCoV-19/Wales/QEUH-9FB63A/2020\|EPI_ISL_600594\|2020 | ···················· | ···················· | ···················· | ···················· | ···················· | ···················· |
| N | 1596 | hCoV-19/Scotland/QEUH-9FA3CB/2020\|EPI_ISL_600598\|2 | ···················· | ···················· | ···················· | ···················· | ···················· | ···················· |
| N | 1597 | hCoV-19/Scotland/QEUH-9FB3F7/2020\|EPI_ISL_600599\|2 | ···················· | ···················· | ···················· | ···················· | ···················· | ···················· |
| N | 1598 | hCoV-19/England/CAMC-9D1EE7/2020\|EPI_ISL_601700\|20 | ···················· | ···················· | ···················· | ···················· | ···················· | ···················· |
| N | 1599 | hCoV-19/England/MILK-9D4822/2020\|EPI_ISL_601701\|20 | ···················· | ···················· | ···················· | ···················· | ···················· | ···················· |
| N | 1600 | hCoV-19/England/MILK-9D48AA/2020\|EPI_ISL_601702\|20 | ···················· | ···················· | ···················· | ···················· | ···················· | ···················· |
| N | 1601 | hCoV-19/Wales/MILK-9D4CF9/2020\|EPI_ISL_601703\|2020 | ···················· | ···················· | ···················· | ···················· | ···················· | ···················· |
| N | 1602 | hCoV-19/Wales/MILK-9D4CDB/2020\|EPI_ISL_601705\|2020 | ···················· | ···················· | ···················· | ···················· | ···················· | ···················· |
| N | 1603 | hCoV-19/England/MILK-9D479E/2020\|EPI_ISL_601706\|20 | ···················· | ·A·················· | ···················· | ···················· | ···················· | ··A················· |
| N | 1604 | hCoV-19/Wales/MILK-9D3B2A/2020\|EPI_ISL_601707\|2020 | ···················· | ···················· | ···················· | ···················· | ···················· | ···················· |
| N | 1605 | hCoV-19/England/MILK-9D4716/2020\|EPI_ISL_601708\|20 | ···················· | ···················· | ···················· | ···················· | ···················· | ···················· |
| N | 1606 | hCoV-19/England/MILK-9D4E6C/2020\|EPI_ISL_601710\|20 | ···················· | ···················· | ···················· | ···················· | ···················· | ···················· |
| N | 1607 | hCoV-19/England/MILK-9D48D7/2020\|EPI_ISL_601711\|20 | ···················· | ···················· | ···················· | ···················· | ···················· | ···················· |
| N | 1608 | hCoV-19/England/MILK-9D44A6/2020\|EPI_ISL_601712\|20 | ···················· | ···················· | ···················· | ···················· | ···················· | ···················· |
| N | 1609 | hCoV-19/England/MILK-9D3EF4/2020\|EPI_ISL_601713\|20 | ···················· | ···················· | ···················· | ···················· | ···················· | ···················· |
| N | 1610 | hCoV-19/Wales/MILK-9D494D/2020\|EPI_ISL_601714\|2020 | ···················· | ···················· | ···················· | ···················· | ···················· | ···················· |
| N | 1611 | hCoV-19/England/CAMC-9D1D17/2020\|EPI_ISL_601715\|20 | ···················· | ·A·················· | ···················· | ···················· | ···················· | ··A················· |
| N | 1612 | hCoV-19/Wales/MILK-9D4E7B/2020\|EPI_ISL_601716\|2020 | ···················· | ···················· | ···················· | ···················· | ···················· | ···················· |
| N | 1613 | hCoV-19/England/MILK-9D4B74/2020\|EPI_ISL_601717\|20 | ···················· | ···················· | ···················· | ···················· | ···················· | ···················· |
| N | 1614 | hCoV-19/England/MILK-9D3F1F/2020\|EPI_ISL_601718\|20 | ···················· | ···················· | ···················· | ···················· | ···················· | ···················· |
| N | 1615 | hCoV-19/Wales/MILK-9D3C09/2020\|EPI_ISL_601719\|2020 | ···················· | ···················· | ···················· | ···················· | ···················· | ···················· |
| N | 1616 | hCoV-19/England/MILK-9D40A2/2020\|EPI_ISL_601720\|20 | ···················· | ···················· | ···················· | ···················· | ···················· | ···················· |
| N | 1617 | hCoV-19/England/MILK-9D3EE5/2020\|EPI_ISL_601722\|20 | ···················· | ···················· | ···················· | ···················· | ···················· | ···················· |
| N | 1618 | hCoV-19/England/MILK-9D47AD/2020\|EPI_ISL_601723\|20 | ···················· | ···················· | ···················· | ···················· | ···················· | ···················· |
| N | 1619 | hCoV-19/England/CAMC-9D1F7B/2020\|EPI_ISL_601768\|20 | ···················· | ···················· | ···················· | ···················· | ···················· | ···················· |
| N | 1620 | hCoV-19/Wales/MILK-9D4C44/2020\|EPI_ISL_601769\|2020 | ···················· | ···················· | ···················· | ···················· | ···················· | ···················· |
| N | 1621 | hCoV-19/England/CAMC-9D1DAE/2020\|EPI_ISL_601771\|20 | ···················· | ···················· | ···················· | ···················· | ···················· | ···················· |
| N | 1622 | hCoV-19/Wales/MILK-9D3F79/2020\|EPI_ISL_601772\|2020 | ···················· | ···················· | ···················· | ···················· | ···················· | ···················· |
| N | 1623 | hCoV-19/England/MILK-9D45EF/2020\|EPI_ISL_601774\|20 | ···················· | ···················· | ···················· | ···················· | ···················· | ···················· |
| N | 1624 | hCoV-19/Scotland/QEUH-9CFC1F/2020\|EPI_ISL_601845\|2 | ···················· | ···················· | ···················· | ···················· | ···················· | ···················· |
| N | 1625 | hCoV-19/England/QEUH-9D062C/2020\|EPI_ISL_601847\|20 | ···················· | ···················· | ···················· | ···················· | ···················· | ···················· |
| N | 1626 | hCoV-19/Scotland/QEUH-9CFDA3/2020\|EPI_ISL_601848\|2 | ···················· | ···················· | ···················· | ···················· | ···················· | ···················· |
| N | 1627 | hCoV-19/Scotland/QEUH-9D09AB/2020\|EPI_ISL_601849\|2 | ···················· | ·A·················· | ···················· | ···················· | ···················· | ···················· |
| N | 1628 | hCoV-19/England/QEUH-9D05F2/2020\|EPI_ISL_601850\|20 | ···················· | ···················· | ···················· | ···················· | ···················· | ···················· |
| N | 1629 | hCoV-19/Scotland/QEUH-9D08AE/2020\|EPI_ISL_601851\|2 | ···················· | ···················· | ···················· | ···················· | ···················· | ···················· |
| N | 1630 | hCoV-19/Scotland/QEUH-9D11C0/2020\|EPI_ISL_601852\|2 | ···················· | ···················· | ···················· | ···················· | ···················· | ···················· |
| N | 1631 | hCoV-19/Scotland/QEUH-9D0CEE/2020\|EPI_ISL_601853\|2 | ···················· | ···················· | ···················· | ···················· | ···················· | ···················· |
| N | 1632 | hCoV-19/Scotland/QEUH-9CFEA0/2020\|EPI_ISL_601854\|2 | ···················· | ···················· | ···················· | ···················· | ···················· | ···················· |
| N | 1633 | hCoV-19/Scotland/QEUH-9D096F/2020\|EPI_ISL_601855\|2 | ···················· | ···················· | ···················· | ···················· | ···················· | ···················· |
| N | 1634 | hCoV-19/Scotland/QEUH-9D0A7B/2020\|EPI_ISL_601858\|2 | ···················· | ···················· | ···················· | ···················· | ···················· | ···················· |
| N | 1635 | hCoV-19/Scotland/QEUH-9D0E24/2020\|EPI_ISL_601859\|2 | ···················· | ···················· | ···················· | ···················· | ···················· | ···················· |
| N | 1636 | hCoV-19/Scotland/QEUH-9D07FC/2020\|EPI_ISL_601860\|2 | ···················· | ···················· | ···················· | ···················· | ···················· | ···················· |
| N | 1637 | hCoV-19/Scotland/QEUH-9D09C9/2020\|EPI_ISL_601861\|2 | ···················· | ···················· | ···················· | ···················· | ···················· | ···················· |
| N | 1638 | hCoV-19/Scotland/QEUH-9D0CFD/2020\|EPI_ISL_601862\|2 | ···················· | ···················· | ···················· | ···················· | ···················· | ···················· |
| N | 1639 | hCoV-19/Scotland/QEUH-9D097E/2020\|EPI_ISL_601863\|2 | ···················· | ···················· | ···················· | ···················· | ···················· | ···················· |
| N | 1640 | hCoV-19/Scotland/QEUH-9D0E06/2020\|EPI_ISL_601865\|2 | ···················· | ···················· | ···················· | ···················· | ···················· | ···················· |
| N | 1641 | hCoV-19/Scotland/QEUH-9CFF16/2020\|EPI_ISL_601866\|2 | ···················· | ···················· | ···················· | ···················· | ···················· | ···················· |
| N | 1642 | hCoV-19/Scotland/QEUH-9D06B3/2020\|EPI_ISL_601867\|2 | ···················· | ·A·················· | ···················· | ···················· | ···················· | ··A················· |
| N | 1643 | hCoV-19/Scotland/QEUH-9CE5F7/2020\|EPI_ISL_601868\|2 | ···················· | ···················· | ···················· | ···················· | ···················· | ···················· |
| N | 1644 | hCoV-19/Scotland/QEUH-9CF365/2020\|EPI_ISL_601869\|2 | ···················· | ···················· | ···················· | ···················· | ···················· | ···················· |
| N | 1645 | hCoV-19/Scotland/QEUH-9CE6C7/2020\|EPI_ISL_601870\|2 | ···················· | ···················· | ···················· | ···················· | ···················· | ···················· |
| N | 1646 | hCoV-19/Scotland/QEUH-9CE384/2020\|EPI_ISL_601871\|2 | ···················· | ···················· | ···················· | ···················· | ···················· | ···················· |
| N | 1647 | hCoV-19/Scotland/QEUH-9CF8C0/2020\|EPI_ISL_601872\|2 | ···················· | ···················· | ···················· | ···················· | ···················· | ···················· |
| N | 1648 | hCoV-19/Scotland/QEUH-9CE25A/2020\|EPI_ISL_601873\|2 | ···················· | ···················· | ···················· | ···················· | ···················· | ···················· |
| N | 1649 | hCoV-19/Scotland/QEUH-9CEAAD/2020\|EPI_ISL_601874\|2 | ···················· | ·A·················· | ···················· | ···················· | ···················· | ···················· |
| N | 1650 | hCoV-19/Scotland/QEUH-9CE982/2020\|EPI_ISL_601875\|2 | ···················· | ···················· | ···················· | ···················· | ···················· | ···················· |
| N | 1651 | hCoV-19/Scotland/QEUH-9CE3A2/2020\|EPI_ISL_601876\|2 | ···················· | ···················· | ···················· | ···················· | ···················· | ···················· |
| N | 1652 | hCoV-19/Scotland/QEUH-9CE7D3/2020\|EPI_ISL_601877\|2 | ···················· | ···················· | ···················· | ···················· | ···················· | ···················· |
| N | 1653 | hCoV-19/Scotland/QEUH-9CE797/2020\|EPI_ISL_601878\|2 | ···················· | ···················· | ···················· | ···················· | ···················· | ···················· |
| N | 1654 | hCoV-19/Scotland/QEUH-9CE964/2020\|EPI_ISL_601879\|2 | ···················· | ···················· | ···················· | ···················· | ···················· | ···················· |
| N | 1655 | hCoV-19/Scotland/QEUH-9CF8B1/2020\|EPI_ISL_601880\|2 | ···················· | ···················· | ···················· | ···················· | ·····T·············· | ···················· |
| N | 1656 | hCoV-19/Scotland/QEUH-9CE3B1/2020\|EPI_ISL_601882\|2 | ···················· | ···················· | ···················· | ···················· | ···················· | ···················· |
| N | 1657 | hCoV-19/Scotland/QEUH-9CF796/2020\|EPI_ISL_601883\|2 | ···················· | ···················· | ···················· | ···················· | ···················· | ···················· |
| N | 1658 | hCoV-19/Scotland/QEUH-9CF338/2020\|EPI_ISL_601884\|2 | ···················· | ···················· | ···················· | ···················· | ···················· | ···················· |
| N | 1659 | hCoV-19/Scotland/QEUH-9CE67C/2020\|EPI_ISL_601885\|2 | ···················· | ···················· | ···················· | ···················· | ···················· | ···················· |
| N | 1660 | hCoV-19/Scotland/QEUH-9CF7A5/2020\|EPI_ISL_601886\|2 | ···················· | ···················· | ···················· | ···················· | ···················· | ···················· |
| N | 1661 | hCoV-19/Scotland/QEUH-9CE68B/2020\|EPI_ISL_601887\|2 | ···················· | ···················· | ···················· | ···················· | ···················· | ···················· |
| N | 1662 | hCoV-19/Scotland/QEUH-9CF022/2020\|EPI_ISL_601888\|2 | ···················· | ···················· | ···················· | ···················· | ···················· | ···················· |
| N | 1663 | hCoV-19/USA/MN-QDX-1654/2020\|EPI_ISL_604000\|2020-0 | ···················· | ···················· | ···················· | ···················· | ···················· | ···················· |
| N | 1664 | hCoV-19/USA/MN-QDX-1659/2020\|EPI_ISL_604001\|2020-0 | ···················· | ···················· | ···················· | ···················· | ···················· | ···················· |
| N | 1665 | hCoV-19/USA/MN-QDX-1663/2020\|EPI_ISL_604002\|2020-0 | ···················· | ···················· | ···················· | ···················· | ···················· | ···················· |
| N | 1666 | hCoV-19/USA/MN-QDX-1664/2020\|EPI_ISL_604003\|2020-0 | ···················· | ···················· | ···················· | ···················· | ···················· | ···················· |
| N | 1667 | hCoV-19/USA/MN-QDX-1666/2020\|EPI_ISL_604004\|2020-0 | ···················· | ···················· | ···················· | ···················· | ···················· | ···················· |

| | | | | | | | | |
|---|---|---|---|---|---|---|---|---|
| N | 1668 | hCoV-19/USA/MN-QDX-1673/2020\|EPI_ISL_604005\|2020-0 | ···················· | ···················· | ···················· | ···················· | ···················· | ···················· |
| N | 1669 | hCoV-19/USA/MN-QDX-1688/2020\|EPI_ISL_604006\|2020-0 | ···················· | ···················· | ···················· | ···················· | ···················· | ···················· |
| N | 1670 | hCoV-19/USA/MN-QDX-1691/2020\|EPI_ISL_604007\|2020-0 | ···················· | ···················· | ···················· | ···················· | ···················· | ···················· |
| N | 1671 | hCoV-19/USA/MN-QDX-1694/2020\|EPI_ISL_604008\|2020-0 | ···················· | ···················· | ···················· | ···················· | ···················· | ···················· |
| N | 1672 | hCoV-19/USA/MN-QDX-1695/2020\|EPI_ISL_604009\|2020-0 | ···················· | ···················· | ···················· | ···················· | ···················· | ···················· |
| N | 1673 | hCoV-19/USA/MN-QDX-1697/2020\|EPI_ISL_604010\|2020-0 | ···················· | ···················· | ···················· | ···················· | ···················· | ···················· |
| N | 1674 | hCoV-19/USA/MN-QDX-1698/2020\|EPI_ISL_604011\|2020-0 | ···················· | ···················· | ···················· | ···················· | ···················· | ···················· |
| N | 1675 | hCoV-19/USA/OR-QDX-1634/2020\|EPI_ISL_604012\|2020-0 | ···················· | ···················· | ···················· | ···················· | ···················· | ···················· |
| N | 1676 | hCoV-19/USA/WA-QDX-1648/2020\|EPI_ISL_604013\|2020-0 | ···················· | ···················· | ···················· | ···················· | ···················· | ···················· |
| N | 1677 | hCoV-19/USA/AZ-QDX-1709/2020\|EPI_ISL_604014\|2020-0 | ···················· | ···················· | ···················· | ···················· | ···················· | ···················· |
| N | 1678 | hCoV-19/USA/AZ-QDX-1729/2020\|EPI_ISL_604015\|2020-0 | ···················· | ···················· | ···················· | ···················· | ···················· | ···················· |
| N | 1679 | hCoV-19/USA/AZ-QDX-1765/2020\|EPI_ISL_604016\|2020-0 | ···················· | ···················· | ···················· | ···················· | ···················· | ···················· |
| N | 1680 | hCoV-19/USA/CA-QDX-1705/2020\|EPI_ISL_604017\|2020-0 | ···················· | ···················· | ···················· | ···················· | ···················· | ···················· |
| N | 1681 | hCoV-19/USA/CA-QDX-1713/2020\|EPI_ISL_604018\|2020-0 | ···················· | ···················· | ···················· | ···················· | ···················· | ···················· |
| N | 1682 | hCoV-19/USA/CA-QDX-1758/2020\|EPI_ISL_604019\|2020-0 | ···················· | ···················· | ···················· | ···················· | ···················· | ···················· |
| N | 1683 | hCoV-19/USA/CA-QDX-1773/2020\|EPI_ISL_604020\|2020-0 | ···················· | ···················· | ···················· | ···················· | ···················· | ···················· |
| N | 1684 | hCoV-19/USA/CT-QDX-1735/2020\|EPI_ISL_604021\|2020-0 | ···················· | ···················· | ···················· | ···················· | ···················· | ···················· |
| N | 1685 | hCoV-19/USA/FL-QDX-1707/2020\|EPI_ISL_604022\|2020-0 | ···················· | ···················· | ···················· | ···················· | ···················· | ···················· |
| N | 1686 | hCoV-19/USA/FL-QDX-1775/2020\|EPI_ISL_604023\|2020-0 | ···················· | ···················· | ···················· | ···················· | ···················· | ···················· |
| N | 1687 | hCoV-19/USA/GA-QDX-1717/2020\|EPI_ISL_604024\|2020-0 | ···················· | ···················· | ···················· | ···················· | ···················· | ···················· |
| N | 1688 | hCoV-19/USA/GA-QDX-1742/2020\|EPI_ISL_604025\|2020-0 | ···················· | ···················· | ···················· | ···················· | ···················· | ···················· |
| N | 1689 | hCoV-19/USA/GA-QDX-1762/2020\|EPI_ISL_604026\|2020-0 | ···················· | ···················· | ···················· | ···················· | ···················· | ···················· |
| N | 1690 | hCoV-19/USA/GA-QDX-1771/2020\|EPI_ISL_604027\|2020-0 | ···················· | ···················· | ···················· | ···················· | ···················· | ···················· |
| N | 1691 | hCoV-19/USA/ID-QDX-1719/2020\|EPI_ISL_604028\|2020-0 | ···················· | ···················· | ···················· | ···················· | ···················· | ···················· |
| N | 1692 | hCoV-19/USA/ID-QDX-1720/2020\|EPI_ISL_604029\|2020-0 | ···················· | ···················· | ···················· | ···················· | ···················· | ···················· |
| N | 1693 | hCoV-19/USA/ID-QDX-1725/2020\|EPI_ISL_604030\|2020-0 | ···················· | ···················· | ···················· | ···················· | ···················· | ···················· |
| N | 1694 | hCoV-19/USA/ID-QDX-1727/2020\|EPI_ISL_604031\|2020-0 | ···················· | ···················· | ···················· | ···················· | ···················· | ···················· |
| N | 1695 | hCoV-19/USA/ID-QDX-1749/2020\|EPI_ISL_604032\|2020-0 | ···················· | ···················· | ···················· | ···················· | ···················· | ···················· |
| N | 1696 | hCoV-19/USA/ID-QDX-1756/2020\|EPI_ISL_604033\|2020-0 | ···················· | ···················· | ···················· | ···················· | ···················· | ···················· |
| N | 1697 | hCoV-19/USA/ID-QDX-1776/2020\|EPI_ISL_604034\|2020-0 | ···················· | ···················· | ···················· | ···················· | ···················· | ···················· |
| N | 1698 | hCoV-19/USA/ID-QDX-1777/2020\|EPI_ISL_604035\|2020-0 | ···················· | ···················· | ···················· | ···················· | ···················· | ···················· |
| N | 1699 | hCoV-19/USA/IN-QDX-1731/2020\|EPI_ISL_604036\|2020-0 | ···················· | ···················· | ···················· | ···················· | ···················· | ···················· |
| N | 1700 | hCoV-19/USA/ME-QDX-1745/2020\|EPI_ISL_604037\|2020-0 | ···················· | ···················· | ···················· | ···················· | ···················· | ···················· |
| N | 1701 | hCoV-19/USA/MI-QDX-1703/2020\|EPI_ISL_604038\|2020-0 | ···················· | ···················· | ···················· | ···················· | ···················· | ···················· |
| N | 1702 | hCoV-19/USA/MI-QDX-1704/2020\|EPI_ISL_604039\|2020-0 | ···················· | ···················· | ···················· | ···················· | ···················· | ···················· |
| N | 1703 | hCoV-19/USA/UT-UPHL-2009573/2020\|EPI_ISL_605251\|20 | ···················· | ·A·················· | ···················· | ···················· | ···················· | ···················· |
| N | 1704 | hCoV-19/USA/UT-UPHL-2009574/2020\|EPI_ISL_605252\|20 | ···················· | ···················· | ···················· | ···················· | ···················· | ···················· |
| N | 1705 | hCoV-19/USA/UT-UPHL-2009576/2020\|EPI_ISL_605254\|20 | ···················· | ···················· | ···················· | ···················· | ···················· | ···················· |
| N | 1706 | hCoV-19/USA/UT-UPHL-2009577/2020\|EPI_ISL_605255\|20 | ···················· | ···················· | ···················· | ···················· | ···················· | ···················· |
| N | 1707 | hCoV-19/USA/UT-UPHL-2009578/2020\|EPI_ISL_605256\|20 | ···················· | ·A·················· | ···················· | ···················· | ···················· | ···················· |
| N | 1708 | hCoV-19/USA/UT-UPHL-2009579/2020\|EPI_ISL_605257\|20 | ···················· | ···················· | ···················· | ···················· | ···················· | ···················· |
| N | 1709 | hCoV-19/USA/UT-UPHL-2009580/2020\|EPI_ISL_605258\|20 | ···················· | ···················· | ···················· | ···················· | ···················· | ···················· |
| N | 1710 | hCoV-19/USA/UT-UPHL-2009581/2020\|EPI_ISL_605259\|20 | ···················· | ···················· | ···················· | ···················· | ···················· | ···················· |
| N | 1711 | hCoV-19/USA/UT-UPHL-2009583/2020\|EPI_ISL_605261\|20 | ···················· | ···················· | ···················· | ···················· | ···················· | ···················· |
| N | 1712 | hCoV-19/USA/UT-UPHL-2009584/2020\|EPI_ISL_605262\|20 | ···················· | ·A·················· | ···················· | ···················· | ···················· | ···················· |
| N | 1713 | hCoV-19/USA/UT-UPHL-2009585/2020\|EPI_ISL_605263\|20 | ···················· | ·A·················· | ···················· | ···················· | ···················· | ···················· |
| N | 1714 | hCoV-19/USA/UT-UPHL-2009586/2020\|EPI_ISL_605264\|20 | ···················· | ···················· | ···················· | ···················· | ···················· | ···················· |
| N | 1715 | hCoV-19/USA/UT-UPHL-2009588/2020\|EPI_ISL_605266\|20 | ···················· | ···················· | ···················· | ···················· | ···················· | ···················· |
| N | 1716 | hCoV-19/USA/UT-UPHL-2009590/2020\|EPI_ISL_605268\|20 | ···················· | ···················· | ···················· | ···················· | ···················· | ···················· |
| N | 1717 | hCoV-19/USA/UT-UPHL-2009591/2020\|EPI_ISL_605269\|20 | ···················· | ···················· | ···················· | ···················· | ···················· | ···················· |
| N | 1718 | hCoV-19/USA/UT-UPHL-2009592/2020\|EPI_ISL_605270\|20 | ···················· | ···················· | ···················· | ···················· | ···················· | ···················· |
| N | 1719 | hCoV-19/USA/UT-UPHL-2009593/2020\|EPI_ISL_605271\|20 | ···················· | ···················· | ···················· | ···················· | ···················· | ···················· |
| N | 1720 | hCoV-19/USA/UT-UPHL-2009595/2020\|EPI_ISL_605273\|20 | ···················· | ·A·················· | ···················· | ···················· | ···················· | ···················· |
| N | 1721 | hCoV-19/USA/UT-UPHL-2009596/2020\|EPI_ISL_605274\|20 | ···················· | ···················· | ···················· | ···················· | ···················· | ···················· |
| N | 1722 | hCoV-19/USA/UT-UPHL-2009597/2020\|EPI_ISL_605275\|20 | ···················· | ···················· | ···················· | ···················· | ···················· | ···················· |
| N | 1723 | hCoV-19/USA/UT-UPHL-2009600/2020\|EPI_ISL_605278\|20 | ···················· | ·A·················· | ···················· | ···················· | ···················· | ···················· |
| N | 1724 | hCoV-19/USA/UT-UPHL-2009601/2020\|EPI_ISL_605279\|20 | ···················· | ···················· | ···················· | ···················· | ···················· | ···················· |
| N | 1725 | hCoV-19/USA/UT-UPHL-2009602/2020\|EPI_ISL_605280\|20 | ···················· | ···················· | ···················· | ···················· | ···················· | ···················· |
| N | 1726 | hCoV-19/USA/UT-UPHL-2009603/2020\|EPI_ISL_605281\|20 | ···················· | ···················· | ···················· | ···················· | ···················· | ···················· |
| N | 1727 | hCoV-19/USA/UT-UPHL-2009604/2020\|EPI_ISL_605282\|20 | ···················· | ···················· | ···················· | ···················· | ···················· | ···················· |
| N | 1728 | hCoV-19/USA/UT-UPHL-2009605/2020\|EPI_ISL_605283\|20 | ···················· | ···················· | ···················· | ···················· | ···················· | ···················· |
| N | 1729 | hCoV-19/USA/UT-UPHL-2009606/2020\|EPI_ISL_605284\|20 | ···················· | ···················· | ···················· | ···················· | ···················· | ···················· |
| N | 1730 | hCoV-19/USA/UT-UPHL-2009607/2020\|EPI_ISL_605285\|20 | ···················· | ·A·················· | ···················· | ···················· | ···················· | ···················· |
| N | 1731 | hCoV-19/USA/UT-UPHL-2009608/2020\|EPI_ISL_605286\|20 | ···················· | ···················· | ···················· | ···················· | ···················· | ···················· |
| N | 1732 | hCoV-19/USA/UT-UPHL-2009609/2020\|EPI_ISL_605287\|20 | ···················· | ···················· | ···················· | ···················· | ···················· | ···················· |
| N | 1733 | hCoV-19/USA/UT-UPHL-2009610/2020\|EPI_ISL_605288\|20 | ···················· | ···················· | ···················· | ···················· | ···················· | ···················· |
| N | 1734 | hCoV-19/USA/UT-UPHL-2009612/2020\|EPI_ISL_605290\|20 | ···················· | ···················· | ···················· | ···················· | ···················· | ···················· |
| N | 1735 | hCoV-19/USA/UT-UPHL-2009613/2020\|EPI_ISL_605291\|20 | ···················· | ···················· | ···················· | ···················· | ···················· | ···················· |
| N | 1736 | hCoV-19/USA/UT-UPHL-2009614/2020\|EPI_ISL_605292\|20 | ···················· | ···················· | ···················· | ···················· | ···················· | ···················· |
| N | 1737 | hCoV-19/USA/UT-UPHL-2009615/2020\|EPI_ISL_605293\|20 | ···················· | ···················· | ···················· | ···················· | ···················· | ···················· |
| N | 1738 | hCoV-19/USA/UT-UPHL-2009616/2020\|EPI_ISL_605294\|20 | ···················· | ·A·················· | ···················· | ···················· | ···················· | ···················· |
| N | 1739 | hCoV-19/USA/UT-UPHL-2009619/2020\|EPI_ISL_605296\|20 | ···················· | ···················· | ···················· | ···················· | ···················· | ···················· |
| N | 1740 | hCoV-19/USA/UT-UPHL-2009620/2020\|EPI_ISL_605297\|20 | ···················· | ···················· | ···················· | ···················· | ···················· | ···················· |
| N | 1741 | hCoV-19/USA/UT-UPHL-2009621/2020\|EPI_ISL_605298\|20 | ···················· | ···················· | ···················· | ···················· | ···················· | ···················· |
| N | 1742 | hCoV-19/USA/UT-UPHL-2009623/2020\|EPI_ISL_605300\|20 | ···················· | ···················· | ···················· | ···················· | ···················· | ···················· |
| N | 1743 | hCoV-19/USA/UT-UPHL-2009624/2020\|EPI_ISL_605301\|20 | ···················· | ···················· | ···················· | ···················· | ·········A·········· | ···················· |
| N | 1744 | hCoV-19/USA/UT-UPHL-2009625/2020\|EPI_ISL_605302\|20 | ···················· | ···················· | ···················· | ···················· | ···················· | ···················· |
| N | 1745 | hCoV-19/USA/UT-UPHL-2009628/2020\|EPI_ISL_605304\|20 | ···················· | ···················· | ···················· | ···················· | ···················· | ···················· |
| N | 1746 | hCoV-19/USA/UT-UPHL-2009631/2020\|EPI_ISL_605307\|20 | ···················· | ···················· | ···················· | ···················· | ···················· | ···················· |
| N | 1747 | hCoV-19/USA/UT-UPHL-2009632/2020\|EPI_ISL_605308\|20 | ···················· | ···················· | ···················· | ···················· | ···················· | ···················· |

| | | | | | | | |
|---|---|---|---|---|---|---|---|
| N | 1748 hCoV-19/USA/UT-UPHL-2009633/2020\|EPI_ISL_605309\|20 | ···················· | ···················· | ···················· | ···················· | ···················· | ···················· |
| N | 1749 hCoV-19/USA/UT-UPHL-2009634/2020\|EPI_ISL_605310\|20 | ···················· | ···················· | ···················· | ···················· | ···················· | ···················· |
| N | 1750 hCoV-19/USA/UT-UPHL-2009638/2020\|EPI_ISL_605313\|20 | ···················· | ·A·················· | ···················· | ···················· | ···················· | ···················· |
| N | 1751 hCoV-19/USA/UT-UPHL-2009640/2020\|EPI_ISL_605315\|20 | ···················· | ···················· | ···················· | ···················· | ···················· | ···················· |
| N | 1752 hCoV-19/USA/UT-UPHL-2009641/2020\|EPI_ISL_605316\|20 | ···················· | ···················· | ···················· | ···················· | ···················· | ···················· |
| N | 1753 hCoV-19/USA/UT-UPHL-2009642/2020\|EPI_ISL_605317\|20 | ···················· | ···················· | ···················· | ···················· | ···················· | ···················· |
| N | 1754 hCoV-19/USA/UT-UPHL-2009643/2020\|EPI_ISL_605318\|20 | ···················· | ···················· | ···················· | ···················· | ···················· | ···················· |
| N | 1755 hCoV-19/USA/UT-UPHL-2009645/2020\|EPI_ISL_605320\|20 | ···················· | ···················· | ···················· | ···················· | ···················· | ···················· |
| N | 1756 hCoV-19/USA/UT-UPHL-2009646/2020\|EPI_ISL_605321\|20 | ···················· | ···················· | ···················· | ···················· | ···················· | ···················· |
| N | 1757 hCoV-19/USA/UT-UPHL-2009647/2020\|EPI_ISL_605322\|20 | ···················· | ···················· | ···················· | ···················· | ···················· | ···················· |
| N | 1758 hCoV-19/USA/UT-UPHL-2009648/2020\|EPI_ISL_605323\|20 | ···················· | ·A·················· | ···················· | ···················· | ···················· | ···················· |
| N | 1759 hCoV-19/USA/UT-UPHL-2009649/2020\|EPI_ISL_605324\|20 | ···················· | ···················· | ···················· | ···················· | ···················· | ···················· |
| N | 1760 hCoV-19/USA/UT-UPHL-2009650/2020\|EPI_ISL_605325\|20 | ···················· | ···················· | ···················· | ···················· | ···················· | ···················· |
| N | 1761 hCoV-19/USA/UT-UPHL-2009651/2020\|EPI_ISL_605326\|20 | ···················· | ···················· | ···················· | ···················· | ···················· | ···················· |
| N | 1762 hCoV-19/USA/UT-UPHL-2009653/2020\|EPI_ISL_605328\|20 | ···················· | ·A·················· | ···················· | ···················· | ···················· | ···················· |
| N | 1763 hCoV-19/USA/UT-UPHL-2009656/2020\|EPI_ISL_605331\|20 | ···················· | ···················· | ···················· | ···················· | ···················· | ···················· |
| N | 1764 hCoV-19/USA/UT-UPHL-2009657/2020\|EPI_ISL_605332\|20 | ···················· | ···················· | ···················· | ···················· | ···················· | ···················· |
| N | 1765 hCoV-19/USA/UT-UPHL-2009660/2020\|EPI_ISL_605335\|20 | ···················· | ···················· | ···················· | ···················· | ···················· | ···················· |
| N | 1766 hCoV-19/USA/UT-UPHL-2009661/2020\|EPI_ISL_605336\|20 | ···················· | ···················· | ···················· | ···················· | ···················· | ···················· |
| N | 1767 hCoV-19/USA/UT-UPHL-2009662/2020\|EPI_ISL_605337\|20 | ···················· | ·A·················· | ···················· | ···················· | ···················· | ···················· |
| N | 1768 hCoV-19/USA/UT-UPHL-2009663/2020\|EPI_ISL_605338\|20 | ············T······· | ···················· | ···················· | ···················· | ···················· | ···················· |
| N | 1769 hCoV-19/USA/UT-UPHL-2009664/2020\|EPI_ISL_605339\|20 | ···················· | ·A·················· | ···················· | ···················· | ···················· | ···················· |
| N | 1770 hCoV-19/USA/UT-UPHL-2009668/2020\|EPI_ISL_605343\|20 | ···················· | ···················· | ···················· | ···················· | ···················· | ···················· |
| N | 1771 hCoV-19/USA/UT-UPHL-2009669/2020\|EPI_ISL_605344\|20 | ···················· | ···················· | ···················· | ···················· | ···················· | ···················· |
| N | 1772 hCoV-19/USA/UT-UPHL-2009670/2020\|EPI_ISL_605345\|20 | ···········T········ | ···················· | ···················· | ···················· | ···················· | ···················· |
| N | 1773 hCoV-19/USA/UT-UPHL-2009673/2020\|EPI_ISL_605348\|20 | ···················· | ···················· | ···················· | ···················· | ···················· | ···················· |
| N | 1774 hCoV-19/USA/UT-UPHL-2009675/2020\|EPI_ISL_605350\|20 | ···················· | ···················· | ···················· | ···················· | ···················· | ···················· |
| N | 1775 hCoV-19/USA/UT-UPHL-2009676/2020\|EPI_ISL_605351\|20 | ···················· | ···················· | ···················· | ···················· | ···················· | ···················· |
| N | 1776 hCoV-19/USA/UT-UPHL-2009677/2020\|EPI_ISL_605352\|20 | ···················· | ···················· | ···················· | ···················· | ···················· | ···················· |
| N | 1777 hCoV-19/USA/UT-UPHL-2009679/2020\|EPI_ISL_605354\|20 | ···················· | ···················· | ···················· | ···················· | ···················· | ···················· |
| N | 1778 hCoV-19/USA/UT-UPHL-2009680/2020\|EPI_ISL_605355\|20 | ···················· | ···················· | ···················· | ···················· | ···················· | ···················· |
| N | 1779 hCoV-19/USA/UT-UPHL-2009681/2020\|EPI_ISL_605356\|20 | ···················· | ···················· | ···················· | ···················· | ···················· | ···················· |
| N | 1780 hCoV-19/USA/UT-UPHL-2009682/2020\|EPI_ISL_605357\|20 | ···················· | ···················· | ···················· | ···················· | ···················· | ···················· |
| N | 1781 hCoV-19/USA/UT-UPHL-2009683/2020\|EPI_ISL_605358\|20 | ···················· | ·A·················· | ···················· | ···················· | ···················· | ···················· |
| N | 1782 hCoV-19/USA/UT-UPHL-2009684/2020\|EPI_ISL_605359\|20 | ···················· | ···················· | ···················· | ···················· | ···················· | ···················· |
| N | 1783 hCoV-19/USA/UT-UPHL-2009686/2020\|EPI_ISL_605360\|20 | ···················· | ···················· | ···················· | ···················· | ···················· | ···················· |
| N | 1784 hCoV-19/USA/UT-UPHL-2009687/2020\|EPI_ISL_605361\|20 | ···················· | ···················· | ···················· | ···················· | ···················· | ···················· |
| N | 1785 hCoV-19/USA/UT-UPHL-2009688/2020\|EPI_ISL_605362\|20 | ···················· | ···················· | ···················· | ···················· | ········T··········· | ···················· |
| N | 1786 hCoV-19/USA/UT-UPHL-2009689/2020\|EPI_ISL_605363\|20 | ···················· | ···················· | ···················· | ···················· | ···················· | ···················· |
| N | 1787 hCoV-19/USA/UT-UPHL-2009690/2020\|EPI_ISL_605364\|20 | ···················· | ···················· | ···················· | ···················· | ···················· | ···················· |
| N | 1788 hCoV-19/USA/UT-UPHL-2009691/2020\|EPI_ISL_605365\|20 | ···················· | ···················· | ···················· | ···················· | ···················· | ···················· |
| N | 1789 hCoV-19/USA/UT-UPHL-2009692/2020\|EPI_ISL_605366\|20 | ···················· | ···················· | ···················· | ···················· | ···················· | ···················· |
| N | 1790 hCoV-19/USA/UT-UPHL-2009693/2020\|EPI_ISL_605367\|20 | ···················· | ···················· | ···················· | ···················· | ···················· | ···················· |
| N | 1791 hCoV-19/USA/UT-UPHL-2009694/2020\|EPI_ISL_605368\|20 | ···················· | ···················· | ···················· | ···················· | ···················· | ···················· |
| N | 1792 hCoV-19/USA/UT-UPHL-2009695/2020\|EPI_ISL_605369\|20 | ···················· | ·A·················· | ···················· | ···················· | ···················· | ···················· |
| N | 1793 hCoV-19/Scotland/QEUH-9FFE4D/2020\|EPI_ISL_600400\|2 | ···················· | ···················· | ···················· | ···················· | ···················· | ···················· |
| N | 1794 hCoV-19/Scotland/QEUH-9FF672/2020\|EPI_ISL_600401\|2 | ···················· | ···················· | ···················· | ···················· | ···················· | ···················· |
| N | 1795 hCoV-19/England/QEUH-9FFF3B/2020\|EPI_ISL_600402\|20 | ···················· | ···················· | ···················· | ···················· | ···················· | ···················· |
| N | 1796 hCoV-19/England/QEUH-9FF250/2020\|EPI_ISL_600403\|20 | ···················· | ···················· | ···················· | ···················· | ···················· | ···················· |
| N | 1797 hCoV-19/England/QEUH-9FF26F/2020\|EPI_ISL_600404\|20 | ···················· | ···················· | ···················· | ···················· | ···················· | ···················· |
| N | 1798 hCoV-19/England/QEUH-A004EC/2020\|EPI_ISL_600406\|20 | ···················· | ···················· | ···················· | ···················· | ···················· | ···················· |
| N | 1799 hCoV-19/England/QEUH-A004A0/2020\|EPI_ISL_600407\|20 | ···················· | ···················· | ···················· | ···················· | ···················· | ···················· |
| N | 1800 hCoV-19/Scotland/QEUH-9FF681/2020\|EPI_ISL_600408\|2 | ···················· | ···················· | ···················· | ···················· | ···················· | ···················· |
| N | 1801 hCoV-19/England/QEUH-9FF214/2020\|EPI_ISL_600409\|20 | ···················· | ···················· | ···················· | ···················· | ···················· | ···················· |
| N | 1802 hCoV-19/England/QEUH-A00859/2020\|EPI_ISL_600410\|20 | ···················· | ···················· | ···················· | ···················· | ···················· | ···················· |
| N | 1803 hCoV-19/Scotland/QEUH-9FFEE3/2020\|EPI_ISL_600411\|2 | ···················· | ···················· | ···················· | ···················· | ···················· | ···················· |
| N | 1804 hCoV-19/England/QEUH-9FF742/2020\|EPI_ISL_600412\|20 | ···················· | ···················· | ···················· | ···················· | ···················· | ···················· |
| N | 1805 hCoV-19/England/QEUH-9FFDE6/2020\|EPI_ISL_600413\|20 | ···················· | ···················· | ···················· | ···················· | ···················· | ···················· |
| N | 1806 hCoV-19/England/QEUH-A006AA/2020\|EPI_ISL_600415\|20 | ···················· | ···················· | ···················· | ···················· | ···················· | ···················· |
| N | 1807 hCoV-19/England/QEUH-9FF900/2020\|EPI_ISL_600416\|20 | ···················· | ···················· | ···················· | ···················· | ···················· | ···················· |
| N | 1808 hCoV-19/England/QEUH-A008FF/2020\|EPI_ISL_600417\|20 | ···················· | ···················· | ···················· | ···················· | ···················· | ···················· |
| N | 1809 hCoV-19/England/QEUH-9FFBA0/2020\|EPI_ISL_600418\|20 | ···················· | ···················· | ···················· | ···················· | ···················· | ···················· |
| N | 1810 hCoV-19/England/QEUH-9FF496/2020\|EPI_ISL_600419\|20 | ···················· | ···················· | ···················· | ···················· | ···················· | ···················· |
| N | 1811 hCoV-19/England/QEUH-9FF5EE/2020\|EPI_ISL_600420\|20 | ···················· | ·A·················· | ···················· | ···················· | ···················· | ···················· |
| N | 1812 hCoV-19/England/QEUH-9FF95B/2020\|EPI_ISL_600421\|20 | ···················· | ···················· | ···················· | ···················· | ···················· | ···················· |
| N | 1813 hCoV-19/England/QEUH-9FFAEF/2020\|EPI_ISL_600422\|20 | ···················· | ···················· | ···················· | ···················· | ···················· | ···················· |
| N | 1814 hCoV-19/England/QEUH-9FF144/2020\|EPI_ISL_600423\|20 | ···················· | ···················· | ···················· | ···················· | ···················· | ···················· |
| N | 1815 hCoV-19/England/QEUH-9FF89A/2020\|EPI_ISL_600424\|20 | ···················· | ···················· | ···················· | ···················· | ···················· | ···················· |
| N | 1816 hCoV-19/England/QEUH-9FFFFF/2020\|EPI_ISL_600425\|20 | ···················· | ···················· | ···················· | ···················· | ···················· | ···················· |
| N | 1817 hCoV-19/Scotland/QEUH-9FF79D/2020\|EPI_ISL_600426\|2 | ···················· | ···················· | ···················· | ···················· | ···················· | ···················· |
| N | 1818 hCoV-19/England/QEUH-9FFC8F/2020\|EPI_ISL_600427\|20 | ···················· | ···················· | ···················· | ···················· | ···················· | ···················· |
| N | 1819 hCoV-19/England/QEUH-A0022E/2020\|EPI_ISL_600448\|20 | ···················· | ···················· | ···················· | ···················· | ···················· | ···················· |
| N | 1820 hCoV-19/Scotland/QEUH-9FF690/2020\|EPI_ISL_600450\|2 | ···················· | ···················· | ···················· | ···················· | ···················· | ···················· |
| N | 1821 hCoV-19/England/QEUH-A0025B/2020\|EPI_ISL_600452\|20 | ···················· | ···················· | ···················· | ···················· | ···················· | ···················· |
| N | 1822 hCoV-19/Scotland/QEUH-9FF7CA/2020\|EPI_ISL_600453\|2 | ···················· | ···················· | ···················· | ···················· | ···················· | ···················· |
| N | 1823 hCoV-19/England/QEUH-A001A9/2020\|EPI_ISL_600455\|20 | ···················· | ···················· | ···················· | ···················· | ···················· | ···················· |
| N | 1824 hCoV-19/Scotland/QEUH-9FF7D9/2020\|EPI_ISL_600457\|2 | ···················· | ···················· | ···················· | ···················· | ···················· | ···················· |
| N | 1825 hCoV-19/England/QEUH-A002D3/2020\|EPI_ISL_600458\|20 | ···················· | ···················· | ···················· | ···················· | ···················· | ···················· |
| N | 1826 hCoV-19/England/QEUH-9FF33F/2020\|EPI_ISL_600460\|20 | ···················· | ···················· | ···················· | ···················· | ···················· | ···················· |
| N | 1827 hCoV-19/Scotland/QEUH-9FFEB6/2020\|EPI_ISL_600462\|2 | ···················· | ···················· | ···················· | ···················· | ···················· | ···················· |

| | | | | | | | | |
|---|---|---|---|---|---|---|---|---|
| N | 1828 | hCoV-19/England/QEUH-A000F7/2020\|EPI_ISL_600464\|20 | ···················· | ···················· | ···················· | ···················· | ···················· | ···················· |
| N | 1829 | hCoV-19/Scotland/QEUH-A007D4/2020\|EPI_ISL_600465\|2 | ···················· | ···················· | ···················· | ···················· | ···················· | ···················· |
| N | 1830 | hCoV-19/Scotland/QEUH-9FFE3E/2020\|EPI_ISL_600467\|2 | ···················· | ···················· | ···················· | ···················· | ···················· | ···················· |
| N | 1831 | hCoV-19/Scotland/QEUH-9FF6CD/2020\|EPI_ISL_600469\|2 | ···················· | ···················· | ···················· | ···················· | ···················· | ···················· |
| N | 1832 | hCoV-19/Scotland/QEUH-A003C1/2020\|EPI_ISL_600470\|2 | ···················· | ···················· | ···················· | ···················· | ···················· | ···················· |
| N | 1833 | hCoV-19/England/QEUH-A0074D/2020\|EPI_ISL_600472\|20 | ···················· | ···················· | ···················· | ···················· | ···················· | ···················· |
| N | 1834 | hCoV-19/England/QEUH-9FFBFB/2020\|EPI_ISL_600474\|20 | ···················· | ···················· | ···················· | ···················· | ···················· | ···················· |
| N | 1835 | hCoV-19/England/QEUH-9FF8D6/2020\|EPI_ISL_600475\|20 | ···················· | ···················· | ···················· | ···················· | ···················· | ···················· |
| N | 1836 | hCoV-19/England/QEUH-A0067D/2020\|EPI_ISL_600477\|20 | ···················· | ·A·················· | ···················· | ···················· | ···················· | ··A················· |
| N | 1837 | hCoV-19/Wales/QEUH-A001F4/2020\|EPI_ISL_600479\|2020 | ···················· | ···················· | ···················· | ···················· | ···················· | ···················· |
| N | 1838 | hCoV-19/Wales/MILK-9D4628/2020\|EPI_ISL_601724\|2020 | ···················· | ···················· | ···················· | ···················· | ···················· | ···················· |
| N | 1839 | hCoV-19/England/MILK-9D48B9/2020\|EPI_ISL_601725\|20 | ···················· | ···················· | ···················· | ···················· | ···················· | ···················· |
| N | 1840 | hCoV-19/Wales/MILK-9D3F88/2020\|EPI_ISL_601726\|2020 | ···················· | ···················· | ···················· | ···················· | ···················· | ···················· |
| N | 1841 | hCoV-19/Wales/MILK-9D4C9F/2020\|EPI_ISL_601727\|2020 | ···················· | ···················· | ···················· | ···················· | ···················· | ···················· |
| N | 1842 | hCoV-19/Wales/MILK-9D3C18/2020\|EPI_ISL_601729\|2020 | ···················· | ···················· | ···················· | ···················· | ···················· | ···················· |
| N | 1843 | hCoV-19/England/CAMC-9D2068/2020\|EPI_ISL_601730\|20 | ···················· | ···················· | ···················· | ···················· | ···················· | ···················· |
| N | 1844 | hCoV-19/England/MILK-9D49A7/2020\|EPI_ISL_601732\|20 | ···················· | ···················· | ···················· | ···················· | ···················· | ···················· |
| N | 1845 | hCoV-19/England/CAMC-9D238D/2020\|EPI_ISL_601733\|20 | ···················· | ···················· | ···················· | ···················· | ···················· | ···················· |
| N | 1846 | hCoV-19/England/MILK-9D4BA1/2020\|EPI_ISL_601734\|20 | ···················· | ···················· | ···················· | ···················· | ···················· | ···················· |
| N | 1847 | hCoV-19/Wales/MILK-9D44B5/2020\|EPI_ISL_601735\|2020 | ···················· | ···················· | ···················· | ···················· | ···················· | ···················· |
| N | 1848 | hCoV-19/Wales/MILK-9D3A69/2020\|EPI_ISL_601737\|2020 | ···················· | ···················· | ···················· | ···················· | ···················· | ···················· |
| N | 1849 | hCoV-19/England/MILK-9D4910/2020\|EPI_ISL_601738\|20 | ···················· | ·A·················· | ···················· | ···················· | ···················· | ··A················· |
| N | 1850 | hCoV-19/England/MILK-9D4D23/2020\|EPI_ISL_601739\|20 | ···················· | ···················· | ···················· | ···················· | ···················· | ···················· |
| N | 1851 | hCoV-19/England/CAMC-9D1F6C/2020\|EPI_ISL_601740\|20 | ···················· | ·A·················· | ···················· | ···················· | ···················· | ··A················· |
| N | 1852 | hCoV-19/England/MILK-9D3CDC/2020\|EPI_ISL_601741\|20 | ···················· | ···················· | ···················· | ···················· | ···················· | ···················· |
| N | 1853 | hCoV-19/Wales/MILK-9D460A/2020\|EPI_ISL_601742\|2020 | ···················· | ···················· | ···················· | ···················· | ···················· | ···················· |
| N | 1854 | hCoV-19/England/MILK-9D4646/2020\|EPI_ISL_601743\|20 | ···················· | ···················· | ···················· | ···················· | ···················· | ···················· |
| N | 1855 | hCoV-19/England/MILK-9D4BCF/2020\|EPI_ISL_601744\|20 | ···················· | ···················· | ···················· | ···················· | ···················· | ···················· |
| N | 1856 | hCoV-19/England/MILK-9D3E7C/2020\|EPI_ISL_601745\|20 | ···················· | ···················· | ···················· | ···················· | ···················· | ···················· |
| N | 1857 | hCoV-19/Wales/MILK-9D3A87/2020\|EPI_ISL_601746\|2020 | ···················· | ···················· | ···················· | ···················· | ···················· | ···················· |
| N | 1858 | hCoV-19/England/CAMC-9D1D9F/2020\|EPI_ISL_601747\|20 | ···················· | ···················· | ···················· | ···················· | ···················· | ···················· |
| N | 1859 | hCoV-19/England/MILK-9D4A1D/2020\|EPI_ISL_601749\|20 | ···················· | ···················· | ···················· | ···················· | ···················· | ···················· |
| N | 1860 | hCoV-19/Wales/MILK-9D4C08/2020\|EPI_ISL_601750\|2020 | ···················· | ···················· | ···················· | ···················· | ···················· | ···················· |
| N | 1861 | hCoV-19/England/MILK-9D3E03/2020\|EPI_ISL_601752\|20 | ···················· | ···················· | ···················· | ···················· | ···················· | ···················· |
| N | 1862 | hCoV-19/England/MILK-9D48F5/2020\|EPI_ISL_601753\|20 | ···················· | ···················· | ···················· | ···················· | ···················· | ···················· |
| N | 1863 | hCoV-19/Wales/MILK-9D45D0/2020\|EPI_ISL_601754\|2020 | ···················· | ···················· | ···················· | ···················· | ···················· | ···················· |
| N | 1864 | hCoV-19/England/MILK-9D49E3/2020\|EPI_ISL_601757\|20 | ···················· | ···················· | ···················· | ···················· | ···················· | ···················· |
| N | 1865 | hCoV-19/Wales/MILK-9D3BDF/2020\|EPI_ISL_601758\|2020 | ···················· | ···················· | ···················· | ···················· | ···················· | ···················· |
| N | 1866 | hCoV-19/Wales/MILK-9D4EC6/2020\|EPI_ISL_601759\|2020 | ···················· | ···················· | ···················· | ···················· | ···················· | ···················· |
| N | 1867 | hCoV-19/Wales/MILK-9D44C4/2020\|EPI_ISL_601760\|2020 | ···················· | ···················· | ···················· | ···················· | ···················· | ···················· |
| N | 1868 | hCoV-19/England/MILK-9D4576/2020\|EPI_ISL_601761\|20 | ···················· | ···················· | ···················· | ···················· | ···················· | ···················· |
| N | 1869 | hCoV-19/NorthernIreland/MILK-9D4691/2020\|EPI_ISL_6 | ···················· | ···················· | ···················· | ···················· | ···················· | ···················· |
| N | 1870 | hCoV-19/England/MILK-9D48C8/2020\|EPI_ISL_601763\|20 | ···················· | ···················· | ···················· | ···················· | ···T················ | ···················· |
| N | 1871 | hCoV-19/England/CAMC-9D1FB7/2020\|EPI_ISL_601764\|20 | ···················· | ···················· | ···················· | ···················· | ···················· | ···················· |
| N | 1872 | hCoV-19/England/MILK-9D401B/2020\|EPI_ISL_601767\|20 | ···················· | ···················· | ···················· | ···················· | ···················· | ···················· |
| N | 1873 | hCoV-19/Wales/MILK-9D4E99/2020\|EPI_ISL_601770\|2020 | ···················· | ···················· | ···················· | ···················· | ···················· | ···················· |
| N | 1874 | hCoV-19/Wales/MILK-9D4637/2020\|EPI_ISL_601775\|2020 | ···················· | ···················· | ···················· | ···················· | ···················· | ···················· |
| N | 1875 | hCoV-19/England/MILK-9D4707/2020\|EPI_ISL_601776\|20 | ···················· | ···················· | ···················· | ···················· | ···················· | ···················· |
| N | 1876 | hCoV-19/England/MILK-9D49D4/2020\|EPI_ISL_601777\|20 | ···················· | ···················· | ···················· | ···················· | ···················· | ···················· |
| N | 1877 | hCoV-19/England/CAMC-9D22CC/2020\|EPI_ISL_601778\|20 | ···················· | ···················· | ···················· | ···················· | ···················· | ···················· |
| N | 1878 | hCoV-19/England/MILK-9D4D50/2020\|EPI_ISL_601779\|20 | ···················· | ···················· | ···················· | ···················· | ···················· | ···················· |
| N | 1879 | hCoV-19/England/MILK-9D4804/2020\|EPI_ISL_601780\|20 | ···················· | ···················· | ···················· | ···················· | ···················· | ···················· |
| N | 1880 | hCoV-19/England/MILK-9D44D3/2020\|EPI_ISL_601781\|20 | ···················· | ·A·················· | ···················· | ···················· | ···················· | ··A················· |
| N | 1881 | hCoV-19/England/MILK-9D3CBE/2020\|EPI_ISL_601782\|20 | ···················· | ···················· | ···················· | ···················· | ···················· | ···················· |
| N | 1882 | hCoV-19/England/CAMC-9D22BD/2020\|EPI_ISL_601783\|20 | ···················· | ···················· | ···················· | ···················· | ···················· | ···················· |
| N | 1883 | hCoV-19/England/MILK-9D3E8B/2020\|EPI_ISL_601784\|20 | ···················· | ···················· | ···················· | ···················· | ···················· | ···················· |
| N | 1884 | hCoV-19/England/CAMC-9D1EC9/2020\|EPI_ISL_601785\|20 | ···················· | ···················· | ···················· | ···················· | ···················· | ···················· |
| N | 1885 | hCoV-19/Wales/MILK-9D45A3/2020\|EPI_ISL_601786\|2020 | ···················· | ···················· | ···················· | ···················· | ···················· | ···················· |
| N | 1886 | hCoV-19/Scotland/QEUH-9D0774/2020\|EPI_ISL_601787\|2 | ···················· | ···················· | ···················· | ···················· | ···················· | ···················· |
| N | 1887 | hCoV-19/Scotland/QEUH-9D01EF/2020\|EPI_ISL_601788\|2 | ···················· | ···················· | ···················· | ···················· | ···················· | ···················· |
| N | 1888 | hCoV-19/Scotland/QEUH-9CFC3D/2020\|EPI_ISL_601789\|2 | ···················· | ···················· | ···················· | ···················· | ···················· | ···················· |
| N | 1889 | hCoV-19/Scotland/QEUH-9CFECE/2020\|EPI_ISL_601790\|2 | ···················· | ···················· | ···················· | ···················· | ···················· | ···················· |
| N | 1890 | hCoV-19/England/QEUH-9D003D/2020\|EPI_ISL_601791\|20 | ···················· | ···················· | ···················· | ···················· | ···················· | ···················· |
| N | 1891 | hCoV-19/Scotland/QEUH-9D0F5E/2020\|EPI_ISL_601792\|2 | ···················· | ···················· | ···················· | ···················· | ···················· | ···················· |
| N | 1892 | hCoV-19/Scotland/QEUH-9D0CB1/2020\|EPI_ISL_601793\|2 | ···················· | ···················· | ···················· | ···················· | ···················· | ···················· |
| N | 1893 | hCoV-19/Scotland/QEUH-9D0C84/2020\|EPI_ISL_601794\|2 | ···················· | ···················· | ···················· | ···················· | ···················· | ···················· |
| N | 1894 | hCoV-19/Scotland/QEUH-9D0C93/2020\|EPI_ISL_601795\|2 | ···················· | ···················· | ···················· | ···················· | ···················· | ···················· |
| N | 1895 | hCoV-19/England/QEUH-9D0686/2020\|EPI_ISL_601796\|20 | ···················· | ···················· | ···················· | ···················· | ···················· | ···················· |
| N | 1896 | hCoV-19/Scotland/QEUH-9D0E33/2020\|EPI_ISL_601798\|2 | ···················· | ···················· | ···················· | ···················· | ···················· | ···················· |
| N | 1897 | hCoV-19/Scotland/QEUH-9D0440/2020\|EPI_ISL_601799\|2 | ···················· | ···················· | ···················· | ···················· | ···················· | ···················· |
| N | 1898 | hCoV-19/Scotland/QEUH-9B7E8D/2020\|EPI_ISL_602100\|2 | ···················· | ···················· | ···················· | ···················· | ···················· | ···················· |
| N | 1899 | hCoV-19/Scotland/QEUH-9B7C92/2020\|EPI_ISL_602101\|2 | ···················· | ···················· | ···················· | ···················· | ···················· | ···················· |
| N | 1900 | hCoV-19/Scotland/QEUH-9B75D3/2020\|EPI_ISL_602104\|2 | ···················· | ···················· | ···················· | ···················· | ···················· | ···················· |
| N | 1901 | hCoV-19/Scotland/QEUH-9B710C/2020\|EPI_ISL_602105\|2 | ···················· | ···················· | ···················· | ···················· | ···················· | ···················· |
| N | 1902 | hCoV-19/Scotland/QEUH-9B78CB/2020\|EPI_ISL_602107\|2 | ···················· | ···················· | ···················· | ···················· | ···················· | ···················· |
| N | 1903 | hCoV-19/Scotland/QEUH-9B711B/2020\|EPI_ISL_602108\|2 | ···················· | ···················· | ···················· | ···················· | ···················· | ···················· |
| N | 1904 | hCoV-19/Scotland/QEUH-9B76FE/2020\|EPI_ISL_602110\|2 | ···················· | ···················· | ···················· | ···················· | ···················· | ···················· |
| N | 1905 | hCoV-19/Scotland/QEUH-9B7245/2020\|EPI_ISL_602111\|2 | ···················· | ···················· | ···················· | ···················· | ···················· | ···················· |
| N | 1906 | hCoV-19/Scotland/QEUH-9B6E60/2020\|EPI_ISL_602112\|2 | ···················· | ···················· | ···················· | ···················· | ···················· | ···················· |
| N | 1907 | hCoV-19/Scotland/QEUH-9B76EF/2020\|EPI_ISL_602113\|2 | ···················· | ···················· | ···················· | ···················· | ···················· | ···················· |

| | | | | | | | | |
|---|---|---|---|---|---|---|---|---|
| N | 1908 | hCoV-19/Scotland/QEUH-9B77CE/2020\|EPI_ISL_602114\|2 | .................... | .................... | .................... | .................... | .................... | .................... |
| N | 1909 | hCoV-19/Scotland/QEUH-9B7BE0/2020\|EPI_ISL_602115\|2 | .................... | .................... | .................... | .................... | .................... | .................... |
| N | 1910 | hCoV-19/Scotland/QEUH-9B7E9C/2020\|EPI_ISL_602117\|2 | .................... | .................... | .................... | .................... | .................... | .................... |
| N | 1911 | hCoV-19/Scotland/QEUH-9B73CA/2020\|EPI_ISL_602118\|2 | .................... | .................... | .................... | .................... | .................... | .................... |
| N | 1912 | hCoV-19/England/QEUH-9B70A5/2020\|EPI_ISL_602119\|20 | .................... | .................... | .................... | .................... | .................... | .................... |
| N | 1913 | hCoV-19/Scotland/QEUH-9B770A/2020\|EPI_ISL_602120\|2 | .................... | .................... | .................... | .................... | .................... | .................... |
| N | 1914 | hCoV-19/Scotland/QEUH-9B7CB0/2020\|EPI_ISL_602121\|2 | .................... | .................... | .................... | .................... | .................... | .................... |
| N | 1915 | hCoV-19/Scotland/QEUH-9B7F6C/2020\|EPI_ISL_602123\|2 | .................... | ......C............. | .................... | .................... | .................... | .................... |
| N | 1916 | hCoV-19/Scotland/QEUH-9B7CCF/2020\|EPI_ISL_602124\|2 | .................... | .................... | .................... | .................... | .................... | .................... |
| N | 1917 | hCoV-19/Scotland/QEUH-9B6F12/2020\|EPI_ISL_602125\|2 | .................... | .................... | .................... | .................... | .................... | .................... |
| N | 1918 | hCoV-19/Scotland/QEUH-9B7087/2020\|EPI_ISL_602126\|2 | .................... | .................... | .................... | .................... | .................... | .................... |
| N | 1919 | hCoV-19/Scotland/QEUH-9B7737/2020\|EPI_ISL_602127\|2 | .................... | .................... | .................... | .................... | .................... | .................... |
| N | 1920 | hCoV-19/Scotland/QEUH-9B70F0/2020\|EPI_ISL_602129\|2 | .................... | .................... | .................... | .................... | .................... | .................... |
| N | 1921 | hCoV-19/Scotland/QEUH-9B70E1/2020\|EPI_ISL_602130\|2 | .................... | .................... | .................... | .................... | .................... | .................... |
| N | 1922 | hCoV-19/Scotland/QEUH-9B6F9A/2020\|EPI_ISL_602132\|2 | .................... | .................... | .................... | .................... | .................... | .................... |
| N | 1923 | hCoV-19/Scotland/QEUH-9B6FD6/2020\|EPI_ISL_602133\|2 | .................... | .................... | .................... | .................... | .................... | .................... |
| N | 1924 | hCoV-19/Scotland/QEUH-9B7CA1/2020\|EPI_ISL_602134\|2 | .................... | .................... | .................... | .................... | .................... | .................... |
| N | 1925 | hCoV-19/England/QEUH-9B8745/2020\|EPI_ISL_602136\|20 | .................... | .................... | .................... | .................... | .................... | .................... |
| N | 1926 | hCoV-19/England/QEUH-9B8754/2020\|EPI_ISL_602137\|20 | .................... | .................... | .................... | .................... | .................... | .................... |
| N | 1927 | hCoV-19/England/QEUH-9B8833/2020\|EPI_ISL_602138\|20 | .................... | .................... | .................... | .................... | .................... | .................... |
| N | 1928 | hCoV-19/England/QEUH-9B8824/2020\|EPI_ISL_602139\|20 | .................... | .................... | .................... | .................... | .................... | .................... |
| N | 1929 | hCoV-19/Switzerland/GE-ETHZ-300447/2020\|EPI_ISL_60 | .................... | .................... | .................... | .................... | .................... | .................... |
| N | 1930 | hCoV-19/Switzerland/GE-ETHZ-300448/2020\|EPI_ISL_60 | .................... | .................... | .................... | .................... | .................... | .................... |
| N | 1931 | hCoV-19/Switzerland/BE-ETHZ-300449/2020\|EPI_ISL_60 | .................... | .................... | .................... | .................... | .................... | .................... |
| N | 1932 | hCoV-19/Switzerland/ZG-ETHZ-300451/2020\|EPI_ISL_60 | .................... | .................... | .................... | .................... | .................... | .................... |
| N | 1933 | hCoV-19/Switzerland/GR-ETHZ-300452/2020\|EPI_ISL_60 | .................... | .................... | .................... | .................... | ......K............. | .................... |
| N | 1934 | hCoV-19/Switzerland/BE-ETHZ-300453/2020\|EPI_ISL_60 | .................... | .................... | .................... | .................... | .................... | .................... |
| N | 1935 | hCoV-19/Switzerland/BE-ETHZ-300454/2020\|EPI_ISL_60 | .................... | .................... | .................... | .................... | .................... | .................... |
| N | 1936 | hCoV-19/Switzerland/BE-ETHZ-300455/2020\|EPI_ISL_60 | .................... | .................... | .................... | .................... | .................... | .................... |
| N | 1937 | hCoV-19/Switzerland/GR-ETHZ-300457/2020\|EPI_ISL_60 | .................... | .................... | .................... | .................... | ......K............. | .................... |
| N | 1938 | hCoV-19/Switzerland/ZH-ETHZ-300459/2020\|EPI_ISL_60 | .................... | .................... | .................... | .................... | .................... | .................... |
| N | 1939 | hCoV-19/Switzerland/VD-ETHZ-300467/2020\|EPI_ISL_60 | .................... | .................... | .................... | .................... | .................... | .................... |
| N | 1940 | hCoV-19/Switzerland/BL-ETHZ-300468/2020\|EPI_ISL_60 | .................... | .................... | .................... | .................... | .................... | .................... |
| N | 1941 | hCoV-19/Switzerland/VD-ETHZ-300469/2020\|EPI_ISL_60 | .................... | .................... | .................... | .................... | .................... | .................... |
| N | 1942 | hCoV-19/Switzerland/VD-ETHZ-300470/2020\|EPI_ISL_60 | .................... | .................... | .................... | .................... | .................... | .................... |
| N | 1943 | hCoV-19/Switzerland/VD-ETHZ-310355/2020\|EPI_ISL_60 | .................... | .................... | .................... | .................... | .................... | .................... |
| N | 1944 | hCoV-19/Switzerland/VD-ETHZ-310356/2020\|EPI_ISL_60 | .................... | .................... | .................... | .................... | .................... | .................... |
| N | 1945 | hCoV-19/Switzerland/VD-ETHZ-310357/2020\|EPI_ISL_60 | .................... | .................... | .................... | .................... | .................... | .................... |
| N | 1946 | hCoV-19/Switzerland/VD-ETHZ-310358/2020\|EPI_ISL_60 | .................... | .................... | .................... | .................... | .................... | .................... |
| N | 1947 | hCoV-19/Switzerland/VD-ETHZ-310359/2020\|EPI_ISL_60 | .................... | .................... | .................... | .................... | .................... | .................... |
| N | 1948 | hCoV-19/Switzerland/VD-ETHZ-310360/2020\|EPI_ISL_60 | .................... | .................... | .................... | .................... | .................... | .................... |
| N | 1949 | hCoV-19/Switzerland/VD-ETHZ-310362/2020\|EPI_ISL_60 | .................... | .................... | .................... | .................... | .................... | .................... |
| N | 1950 | hCoV-19/Switzerland/VD-ETHZ-310363/2020\|EPI_ISL_60 | .................... | .................... | .................... | .................... | .................... | .................... |
| N | 1951 | hCoV-19/Switzerland/VD-ETHZ-310364/2020\|EPI_ISL_60 | .................... | .................... | .................... | .................... | .................... | .................... |
| N | 1952 | hCoV-19/Switzerland/ZH-ETHZ-310365/2020\|EPI_ISL_60 | .................... | .................... | .................... | .................... | .................... | .................... |
| N | 1953 | hCoV-19/Switzerland/ZH-ETHZ-310366/2020\|EPI_ISL_60 | .................... | .................... | .................... | .................... | .................... | .................... |
| N | 1954 | hCoV-19/Switzerland/ZH-ETHZ-310367/2020\|EPI_ISL_60 | .................... | .................... | .................... | .................... | .................... | .................... |
| N | 1955 | hCoV-19/Switzerland/ZH-ETHZ-310368/2020\|EPI_ISL_60 | .................... | .................... | .................... | .................... | .................... | .................... |
| N | 1956 | hCoV-19/Switzerland/VD-ETHZ-310369/2020\|EPI_ISL_60 | .................... | .................... | .................... | .................... | .................... | .................... |
| N | 1957 | hCoV-19/Switzerland/VD-ETHZ-310370/2020\|EPI_ISL_60 | .................... | .................... | .................... | .................... | .................... | .................... |
| N | 1958 | hCoV-19/Switzerland/VS-ETHZ-310371/2020\|EPI_ISL_60 | .................... | ......A............. | .................... | .................... | .................... | .................... |
| N | 1959 | hCoV-19/Switzerland/VD-ETHZ-310372/2020\|EPI_ISL_60 | .................... | .................... | .................... | .................... | .................... | .................... |
| N | 1960 | hCoV-19/Switzerland/VD-ETHZ-310373/2020\|EPI_ISL_60 | .................... | .................... | .................... | .................... | .................... | .................... |
| N | 1961 | hCoV-19/Switzerland/VS-ETHZ-310374/2020\|EPI_ISL_60 | .................... | ......A............. | .................... | .................... | .................... | .................... |
| N | 1962 | hCoV-19/Switzerland/VD-ETHZ-310375/2020\|EPI_ISL_60 | .................... | .................... | .................... | .................... | .................... | .................... |
| N | 1963 | hCoV-19/Switzerland/VS-ETHZ-310376/2020\|EPI_ISL_60 | .................... | ......A............. | .................... | .................... | .................... | .................... |
| N | 1964 | hCoV-19/Switzerland/VD-ETHZ-310377/2020\|EPI_ISL_60 | .................... | .................... | .................... | .................... | .................... | .................... |
| N | 1965 | hCoV-19/Switzerland/VS-ETHZ-310378/2020\|EPI_ISL_60 | .................... | .................... | .................... | .................... | .................... | .................... |
| N | 1966 | hCoV-19/Switzerland/VD-ETHZ-310379/2020\|EPI_ISL_60 | .................... | .................... | .................... | .................... | .................... | .................... |
| N | 1967 | hCoV-19/Switzerland/VS-ETHZ-310380/2020\|EPI_ISL_60 | .................... | ......A............. | .................... | .................... | .................... | .................... |
| N | 1968 | hCoV-19/Switzerland/BL-ETHZ-310381/2020\|EPI_ISL_60 | .................... | .................... | .................... | .................... | .................... | .................... |
| N | 1969 | hCoV-19/Switzerland/BL-ETHZ-310382/2020\|EPI_ISL_60 | .................... | .................... | .................... | .................... | .................... | .................... |
| N | 1970 | hCoV-19/Switzerland/BL-ETHZ-310383/2020\|EPI_ISL_60 | .................... | .................... | .................... | .................... | .................... | .................... |
| N | 1971 | hCoV-19/Switzerland/SZ-ETHZ-310384/2020\|EPI_ISL_60 | .................... | .................... | .................... | .................... | .................... | .................... |
| N | 1972 | hCoV-19/Switzerland/BL-ETHZ-310385/2020\|EPI_ISL_60 | .................... | .................... | .................... | .................... | .................... | .................... |
| N | 1973 | hCoV-19/Switzerland/BL-ETHZ-310386/2020\|EPI_ISL_60 | .................... | .................... | .................... | .................... | .................... | .................... |
| N | 1974 | hCoV-19/Switzerland/BL-ETHZ-310387/2020\|EPI_ISL_60 | .................... | .................... | .................... | .................... | .................... | .................... |
| N | 1975 | hCoV-19/Switzerland/BL-ETHZ-310388/2020\|EPI_ISL_60 | .................... | .................... | .................... | .................... | .................... | .................... |
| N | 1976 | hCoV-19/Switzerland/SZ-ETHZ-310389/2020\|EPI_ISL_60 | .................... | .................... | .................... | .................... | .................... | .................... |
| N | 1977 | hCoV-19/Switzerland/SZ-ETHZ-310390/2020\|EPI_ISL_60 | .................... | .................... | .................... | .................... | .................... | .................... |
| N | 1978 | hCoV-19/Switzerland/SZ-ETHZ-310391/2020\|EPI_ISL_60 | .................... | .................... | .................... | .................... | .................... | .................... |
| N | 1979 | hCoV-19/Switzerland/NW-ETHZ-310392/2020\|EPI_ISL_60 | .................... | .................... | .................... | .................... | .................... | .................... |
| N | 1980 | hCoV-19/Switzerland/SZ-ETHZ-310394/2020\|EPI_ISL_60 | .................... | .................... | .................... | .................... | .................... | .................... |
| N | 1981 | hCoV-19/Switzerland/AG-ETHZ-310395/2020\|EPI_ISL_60 | .................... | .................... | .................... | .................... | ....K............... | .................... |
| N | 1982 | hCoV-19/USA/NY-QDX-2414/2020\|EPI_ISL_604716\|2020-0 | .................... | .................... | .................... | .................... | .................... | .................... |
| N | 1983 | hCoV-19/USA/NY-QDX-2422/2020\|EPI_ISL_604717\|2020-0 | .................... | .................... | .................... | .................... | .................... | .................... |
| N | 1984 | hCoV-19/USA/OH-QDX-2391/2020\|EPI_ISL_604718\|2020-0 | .................... | .................... | .................... | .................... | .................... | .................... |
| N | 1985 | hCoV-19/USA/PR-QDX-2339/2020\|EPI_ISL_604719\|2020-0 | .................... | .................... | .................... | .................... | .................... | .................... |
| N | 1986 | hCoV-19/USA/TX-QDX-2369/2020\|EPI_ISL_604720\|2020-0 | .................... | .................... | .................... | .................... | .................... | .................... |
| N | 1987 | hCoV-19/USA/TX-QDX-2409/2020\|EPI_ISL_604721\|2020-0 | .................... | .................... | .................... | .................... | .................... | .................... |

N 1988 hCoV-19/USA/UT-QDX-2315/2020|EPI_ISL_604722|2020-0 ···················· ···················· ···················· ···················· ···················· ····················
N 1989 hCoV-19/USA/UT-QDX-2321/2020|EPI_ISL_604723|2020-0 ···················· ···················· ···················· ···················· ···················· ····················
N 1990 hCoV-19/USA/UT-QDX-2347/2020|EPI_ISL_604724|2020-0 ···················· ···················· ···················· ···················· ···················· ····················
N 1991 hCoV-19/USA/UT-QDX-2356/2020|EPI_ISL_604725|2020-0 ···················· ···················· ···················· ···················· ···················· ····················
N 1992 hCoV-19/USA/UT-QDX-2359/2020|EPI_ISL_604726|2020-0 ···················· ···················· ···················· ···················· ···················· ····················
N 1993 hCoV-19/USA/UT-QDX-2407/2020|EPI_ISL_604728|2020-0 ···················· ···················· ···················· ···················· ···················· ····················
N 1994 hCoV-19/USA/UT-QDX-2415/2020|EPI_ISL_604729|2020-0 ···················· ···················· ···················· ···················· ···················· ····················
N 1995 hCoV-19/USA/UT-QDX-2417/2020|EPI_ISL_604730|2020-0 ···················· ···················· ···················· ···················· ···················· ····················
N 1996 hCoV-19/USA/UT-QDX-2423/2020|EPI_ISL_604731|2020-0 ···················· ···················· ···················· ···················· ···················· ····················
N 1997 hCoV-19/USA/WA-QDX-2319/2020|EPI_ISL_604732|2020-0 ···················· ···················· ···················· ···················· ···················· ····················
N 1998 hCoV-19/USA/WA-QDX-2326/2020|EPI_ISL_604733|2020-0 ···················· ···················· ···················· ···················· ···················· ····················
N 1999 hCoV-19/USA/WA-QDX-2327/2020|EPI_ISL_604734|2020-0 ···················· ···················· ···················· ···················· ···················· ····················
N 2000 hCoV-19/USA/WA-QDX-2333/2020|EPI_ISL_604735|2020-0 ···················· ···················· ···················· ···················· ···················· ····················
N 2001 hCoV-19/USA/WA-QDX-2354/2020|EPI_ISL_604736|2020-0 ···················· ···················· ···················· ···················· ···················· ····················
N 2002 hCoV-19/USA/WA-QDX-2376/2020|EPI_ISL_604737|2020-0 ···················· ·············A······ ···················· ···················· ···················· ····················
N 2003 hCoV-19/USA/WI-QDX-2402/2020|EPI_ISL_604738|2020-0 ···················· ···················· ···················· ···················· ···················· ····················
N 2004 hCoV-19/USA/AR-QDX-2472/2020|EPI_ISL_604739|2020-0 ···················· ···················· ···················· ···················· ···················· ····················
N 2005 hCoV-19/USA/CA-QDX-2449/2020|EPI_ISL_604745|2020-0 ···················· ···················· ···················· ···················· ···················· ····················
N 2006 hCoV-19/USA/CA-QDX-2450/2020|EPI_ISL_604746|2020-0 ···················· ···················· ···················· ···················· ···················· ····················
N 2007 hCoV-19/USA/CA-QDX-2457/2020|EPI_ISL_604747|2020-0 ···················· ···················· ···················· ···················· ···················· ····················
N 2008 hCoV-19/USA/CA-QDX-2458/2020|EPI_ISL_604748|2020-0 ···················· ···················· ···················· ···················· ···················· ····················
N 2009 hCoV-19/USA/CA-QDX-2503/2020|EPI_ISL_604760|2020-0 ···················· ···················· ···················· ···················· ···················· ····················
N 2010 hCoV-19/USA/CA-QDX-2507/2020|EPI_ISL_604761|2020-0 ···················· ···················· ···················· ···················· ···················· ····················
N 2011 hCoV-19/USA/CA-QDX-2515/2020|EPI_ISL_604762|2020-0 ···················· ···················· ···················· ···················· ···················· ····················
N 2012 hCoV-19/USA/CA-QDX-2519/2020|EPI_ISL_604763|2020-0 ···················· ···················· ···················· ···················· ···················· ····················
N 2013 hCoV-19/USA/CA-QDX-2520/2020|EPI_ISL_604764|2020-0 ···················· ···················· ···················· ···················· ···················· ····················
N 2014 hCoV-19/USA/CA-QDX-2524/2020|EPI_ISL_604765|2020-0 ···················· ···················· ···················· ···················· ···················· ····················
N 2015 hCoV-19/USA/CA-QDX-2537/2020|EPI_ISL_604766|2020-0 ···················· ···················· ···················· ···················· ···················· ····················
N 2016 hCoV-19/USA/CA-QDX-2540/2020|EPI_ISL_604767|2020-0 ···················· ···················· ···················· ···················· ···················· ····················
N 2017 hCoV-19/USA/CA-QDX-2551/2020|EPI_ISL_604768|2020-0 ···················· ···················· ···················· ···················· ···················· ····················
N 2018 hCoV-19/USA/CA-QDX-2552/2020|EPI_ISL_604769|2020-0 ···················· ···················· ···················· ···················· ···················· ····················
N 2019 hCoV-19/USA/CA-QDX-2555/2020|EPI_ISL_604770|2020-0 ···················· ···················· ···················· ···················· ···················· ····················
N 2020 hCoV-19/USA/CA-QDX-2559/2020|EPI_ISL_604771|2020-0 ···················· ···················· ···················· ···················· ···················· ····················
N 2021 hCoV-19/USA/CA-QDX-2561/2020|EPI_ISL_604772|2020-0 ···················· ···················· ···················· ···················· ···················· ····················
N 2022 hCoV-19/USA/FL-QDX-2456/2020|EPI_ISL_604773|2020-0 ···················· ···················· ···················· ···················· ···················· ····················
N 2023 hCoV-19/USA/FL-QDX-2502/2020|EPI_ISL_604774|2020-0 ···················· ···················· ···················· ···················· ···················· ····················
N 2024 hCoV-19/USA/FL-QDX-2510/2020|EPI_ISL_604775|2020-0 ···················· ···················· ···················· ···················· ···················· ····················
N 2025 hCoV-19/USA/FL-QDX-2513/2020|EPI_ISL_604776|2020-0 ···················· ···················· ···················· ···················· ···················· ····················
N 2026 hCoV-19/USA/GA-QDX-2442/2020|EPI_ISL_604777|2020-0 ···················· ···················· ···················· ···················· ···················· ····················
N 2027 hCoV-19/USA/GA-QDX-2443/2020|EPI_ISL_604778|2020-0 ···················· ···················· ···················· ···················· ···················· ····················
N 2028 hCoV-19/USA/GA-QDX-2461/2020|EPI_ISL_604779|2020-0 ···················· ···················· ···················· ···················· ···················· ····················
N 2029 hCoV-19/USA/GA-QDX-2511/2020|EPI_ISL_604780|2020-0 ···················· ···················· ···················· ···················· ···················· ····················
N 2030 hCoV-19/USA/GA-QDX-2553/2020|EPI_ISL_604781|2020-0 ···················· ···················· ···················· ···················· ···················· ····················
N 2031 hCoV-19/USA/GA-QDX-2558/2020|EPI_ISL_604782|2020-0 ···················· ···················· ···················· ···················· ···················· ····················
N 2032 hCoV-19/USA/MA-QDX-2427/2020|EPI_ISL_604783|2020-0 ···················· ···················· ···················· ···················· ···················· ····················
N 2033 hCoV-19/USA/MO-QDX-2460/2020|EPI_ISL_604784|2020-0 ···················· ···················· ···················· ···················· ···················· ····················
N 2034 hCoV-19/USA/MO-QDX-2516/2020|EPI_ISL_604785|2020-0 ···················· ···················· ···················· ···················· ···················· ····················
N 2035 hCoV-19/USA/NJ-QDX-2430/2020|EPI_ISL_604786|2020-0 ···················· ···················· ···················· ···················· ···················· ····················
N 2036 hCoV-19/USA/NJ-QDX-2432/2020|EPI_ISL_604787|2020-0 ···················· ···················· ···················· ···················· ···················· ····················
N 2037 hCoV-19/USA/NJ-QDX-2468/2020|EPI_ISL_604788|2020-0 ···················· ···················· ···················· ···················· ···················· ····················
N 2038 hCoV-19/USA/NJ-QDX-2495/2020|EPI_ISL_604789|2020-0 ···················· ···················· ···················· ···················· ···················· ····················
N 2039 hCoV-19/USA/NJ-QDX-2518/2020|EPI_ISL_604790|2020-0 ···················· ···················· ···················· ···················· ···················· ····················
N 2040 hCoV-19/USA/NJ-QDX-2541/2020|EPI_ISL_604791|2020-0 ···················· ···················· ···················· ···················· ···················· ····················
N 2041 hCoV-19/USA/NJ-QDX-2544/2020|EPI_ISL_604792|2020-0 ···················· ···················· ···················· ···················· ···················· ····················
N 2042 hCoV-19/USA/NM-QDX-2538/2020|EPI_ISL_604793|2020-0 ···················· ···················· ···················· ···················· ···················· ····················
N 2043 hCoV-19/USA/NY-QDX-2428/2020|EPI_ISL_604794|2020-0 ···················· ···················· ···················· ···················· ···················· ····················
N 2044 hCoV-19/USA/NY-QDX-2431/2020|EPI_ISL_604795|2020-0 ···················· ···················· ···················· ···················· ···················· ····················
N 2045 hCoV-19/USA/NY-QDX-2438/2020|EPI_ISL_604796|2020-0 ···················· ···················· ···················· ···················· ···················· ····················
N 2046 hCoV-19/USA/NY-QDX-2439/2020|EPI_ISL_604797|2020-0 ···················· ···················· ···················· ···················· ···················· ····················
N 2047 hCoV-19/USA/NY-QDX-2444/2020|EPI_ISL_604798|2020-0 ···················· ···················· ···················· ···················· ···················· ····················
N 2048 hCoV-19/USA/NY-QDX-2445/2020|EPI_ISL_604799|2020-0 ···················· ···················· ···················· ···················· ···················· ····················
N 2049 hCoV-19/England/CAMC-A57F7C/2020|EPI_ISL_597039|20 ···················· ···················· ···················· ···················· ···················· ····················
N 2050 hCoV-19/England/CAMC-A589C8/2020|EPI_ISL_597040|20 ···················· ···················· ···················· ···················· ···················· ····················
N 2051 hCoV-19/England/CAMC-A57CFD/2020|EPI_ISL_597041|20 ···················· ···················· ···················· ···················· ···················· ····················
N 2052 hCoV-19/England/CAMC-A58F20/2020|EPI_ISL_597042|20 ···················· ···················· ···················· ···················· ···················· ····················
N 2053 hCoV-19/England/CAMC-A57DAF/2020|EPI_ISL_597043|20 ···················· ···················· ···················· ···················· ···················· ····················
N 2054 hCoV-19/England/CAMC-A58EE7/2020|EPI_ISL_597045|20 ···················· ·A·················· ···················· ···················· ···················· ····················
N 2055 hCoV-19/England/CAMC-A58667/2020|EPI_ISL_597046|20 ···················· ···················· ···················· ···················· ···················· ····················
N 2056 hCoV-19/England/CAMC-A58C29/2020|EPI_ISL_597047|20 ···················· ···················· ···················· ···················· ···················· ····················
N 2057 hCoV-19/England/CAMC-A57E15/2020|EPI_ISL_597048|20 ···················· ···················· ···················· ···················· ···················· ····················
N 2058 hCoV-19/England/CAMC-A57E33/2020|EPI_ISL_597050|20 ···················· ···················· ···················· ···················· ···················· ····················
N 2059 hCoV-19/England/CAMC-A57FB8/2020|EPI_ISL_597051|20 ···················· ···················· ···················· ···················· ···················· ····················
N 2060 hCoV-19/England/CAMC-A5897D/2020|EPI_ISL_597052|20 ···················· ···················· ···················· ···················· ···················· ····················
N 2061 hCoV-19/England/CAMC-A5904A/2020|EPI_ISL_597053|20 ···················· ···················· ···················· ···················· ···················· ····················
N 2062 hCoV-19/England/CAMC-A586A3/2020|EPI_ISL_597054|20 ···················· ·A·················· ···················· ···················· ···················· ···A················
N 2063 hCoV-19/England/CAMC-A58D08/2020|EPI_ISL_597057|20 ···················· ···················· ···················· ···················· ···················· ····················
N 2064 hCoV-19/England/CAMC-A59341/2020|EPI_ISL_597058|20 ···················· ···················· ···················· ···················· ···················· ····················
N 2065 hCoV-19/England/CAMC-A58D17/2020|EPI_ISL_597059|20 ···················· ···················· ···················· ···················· ···················· ····················
N 2066 hCoV-19/England/CAMC-A58B68/2020|EPI_ISL_597060|20 ···················· ···················· ···················· ···················· ···················· ····················
N 2067 hCoV-19/England/CAMC-A58E50/2020|EPI_ISL_597061|20 ···················· ···················· ···················· ···················· ···················· ····················

| | | | | | | | | |
|---|---|---|---|---|---|---|---|---|
| N | 2068 | hCoV-19/England/CAMC-A58807/2020\|EPI_ISL_597062\|20 | ···················· | ···················· | ····················· | ···················· | ···················· | ···················· |
| N | 2069 | hCoV-19/England/CAMC-A58087/2020\|EPI_ISL_597063\|20 | ···················· | ···················· | ····················· | ···················· | ······T············· | ···················· |
| N | 2070 | hCoV-19/England/CAMC-A5812A/2020\|EPI_ISL_597064\|20 | ···················· | ···················· | ····················· | ···················· | ···················· | ···················· |
| N | 2071 | hCoV-19/England/CAMC-A57DEB/2020\|EPI_ISL_597066\|20 | ···················· | ···················· | ····················· | ···················· | ···················· | ···················· |
| N | 2072 | hCoV-19/England/CAMC-A59095/2020\|EPI_ISL_597068\|20 | ···················· | ···················· | ····················· | ···················· | ···················· | ···················· |
| N | 2073 | hCoV-19/England/CAMC-A589E6/2020\|EPI_ISL_597069\|20 | ···················· | ···················· | ····················· | ···················· | ···················· | ···················· |
| N | 2074 | hCoV-19/England/CAMC-A58218/2020\|EPI_ISL_597070\|20 | ···················· | ···················· | ····················· | ···················· | ···················· | ···················· |
| N | 2075 | hCoV-19/England/CAMC-A586B2/2020\|EPI_ISL_597071\|20 | ···················· | ···················· | ····················· | ···················· | ···················· | ···················· |
| N | 2076 | hCoV-19/England/CAMC-A588AD/2020\|EPI_ISL_597072\|20 | ···················· | ···················· | ····················· | ···················· | ···················· | ···················· |
| N | 2077 | hCoV-19/England/CAMC-A59639/2020\|EPI_ISL_597073\|20 | ···················· | ···················· | ····················· | ···················· | ···················· | ···················· |
| N | 2078 | hCoV-19/England/CAMC-A58AB6/2020\|EPI_ISL_597076\|20 | ···················· | ···················· | ····················· | ···················· | ······T············· | ···················· |
| N | 2079 | hCoV-19/England/CAMC-A58DEA/2020\|EPI_ISL_597077\|20 | ···················· | ···················· | ····················· | ···················· | ···················· | ···················· |
| N | 2080 | hCoV-19/England/CAMC-A581DF/2020\|EPI_ISL_597078\|20 | ···················· | ···················· | ····················· | ···················· | ···················· | ···················· |
| N | 2081 | hCoV-19/England/CAMC-A587FB/2020\|EPI_ISL_597079\|20 | ···················· | ···················· | ····················· | ···················· | ···················· | ···················· |
| N | 2082 | hCoV-19/England/CAMC-A58139/2020\|EPI_ISL_597080\|20 | ···················· | ···················· | ····················· | ···················· | ···················· | ···················· |
| N | 2083 | hCoV-19/England/CAMC-A57D18/2020\|EPI_ISL_597081\|20 | ···················· | ·A·················· | ····················· | ···················· | ···················· | ···················· |
| N | 2084 | hCoV-19/England/CAMC-A58BC2/2020\|EPI_ISL_597082\|20 | ···················· | ···················· | ····················· | ···················· | ···················· | ···················· |
| N | 2085 | hCoV-19/England/CAMC-A57C57/2020\|EPI_ISL_597083\|20 | ···················· | ···················· | ····················· | ···················· | ···················· | ···················· |
| N | 2086 | hCoV-19/England/CAMC-A5911A/2020\|EPI_ISL_597084\|20 | ···················· | ···················· | ····················· | ···················· | ···················· | ···················· |
| N | 2087 | hCoV-19/England/CAMC-A58BE0/2020\|EPI_ISL_597085\|20 | ···················· | ·A·················· | ····················· | ···················· | ···················· | ···A················ |
| N | 2088 | hCoV-19/England/CAMC-A581EE/2020\|EPI_ISL_597087\|20 | ···················· | ···················· | ····················· | ···················· | ···················· | ···················· |
| N | 2089 | hCoV-19/England/CAMC-A57D45/2020\|EPI_ISL_597088\|20 | ···················· | ···················· | ····················· | ···················· | ···················· | ···················· |
| N | 2090 | hCoV-19/England/CAMC-A5953C/2020\|EPI_ISL_597089\|20 | ···················· | ···················· | ····················· | ···················· | ···················· | ···················· |
| N | 2091 | hCoV-19/England/CAMC-A58719/2020\|EPI_ISL_597090\|20 | ···················· | ···················· | ····················· | ···················· | ···················· | ···················· |
| N | 2092 | hCoV-19/England/CAMC-A5954B/2020\|EPI_ISL_597091\|20 | ···················· | ···················· | ····················· | ···················· | ···················· | ···················· |
| N | 2093 | hCoV-19/England/CAMC-A58FF3/2020\|EPI_ISL_597092\|20 | ···················· | ···················· | ····················· | ···················· | ···················· | ···················· |
| N | 2094 | hCoV-19/England/CAMC-A592F9/2020\|EPI_ISL_597093\|20 | ···················· | ···················· | ····················· | ···················· | ···················· | ···················· |
| N | 2095 | hCoV-19/England/CAMC-A585E2/2020\|EPI_ISL_597094\|20 | ···················· | ···················· | ····················· | ···················· | ···················· | ···················· |
| N | 2096 | hCoV-19/England/CAMC-A58CED/2020\|EPI_ISL_597095\|20 | ···················· | ···················· | ····················· | ···················· | ···················· | ···················· |
| N | 2097 | hCoV-19/England/CAMC-A59718/2020\|EPI_ISL_597096\|20 | ···················· | ···················· | ····················· | ···················· | ···················· | ···················· |
| N | 2098 | hCoV-19/England/CAMC-A592BD/2020\|EPI_ISL_597097\|20 | ···················· | ···················· | ····················· | ···················· | ···················· | ···················· |
| N | 2099 | hCoV-19/England/CAMC-A59235/2020\|EPI_ISL_597098\|20 | ···················· | ···················· | ····················· | ···················· | ···················· | ···················· |
| N | 2100 | hCoV-19/England/CAMC-A58C74/2020\|EPI_ISL_597100\|20 | ···················· | ·A·················· | ····················· | ···················· | ···················· | ···A················ |
| N | 2101 | hCoV-19/England/CAMC-A59280/2020\|EPI_ISL_597101\|20 | ···················· | ···················· | ····················· | ···················· | ···················· | ···················· |
| N | 2102 | hCoV-19/England/CAMC-A5989D/2020\|EPI_ISL_597102\|20 | ···················· | ···················· | ····················· | ···················· | ···················· | ···················· |
| N | 2103 | hCoV-19/England/CAMC-A596B1/2020\|EPI_ISL_597103\|20 | ···················· | ···················· | ····················· | ···················· | ···················· | ···················· |
| N | 2104 | hCoV-19/England/CAMC-A57CC0/2020\|EPI_ISL_597105\|20 | ···················· | ···················· | ····················· | ···················· | ···················· | ···················· |
| N | 2105 | hCoV-19/England/CAMC-A5939C/2020\|EPI_ISL_597106\|20 | ···················· | ···················· | ····················· | ···················· | ···················· | ···················· |
| N | 2106 | hCoV-19/England/CAMC-A58C0B/2020\|EPI_ISL_597107\|20 | ···················· | ···················· | ····················· | ···················· | ···················· | ···················· |
| N | 2107 | hCoV-19/England/CAMC-A59727/2020\|EPI_ISL_597108\|20 | ···················· | ···················· | ····················· | ···················· | ···················· | ···················· |
| N | 2108 | hCoV-19/England/CAMC-A585F1/2020\|EPI_ISL_597110\|20 | ···················· | ···················· | ····················· | ···················· | ···················· | ···················· |
| N | 2109 | hCoV-19/England/CAMC-A593BA/2020\|EPI_ISL_597111\|20 | ···················· | ···················· | ····················· | ···················· | ···················· | ···················· |
| N | 2110 | hCoV-19/England/CAMC-A583E8/2020\|EPI_ISL_597112\|20 | ···················· | ···················· | ····················· | ···················· | ···················· | ···················· |
| N | 2111 | hCoV-19/England/CAMC-A57F4F/2020\|EPI_ISL_597113\|20 | ···················· | ·A·················· | ····················· | ···················· | ···················· | ···A················ |
| N | 2112 | hCoV-19/England/CAMC-A590C2/2020\|EPI_ISL_597114\|20 | ···················· | ···················· | ····················· | ···················· | ···················· | ···R················ |
| N | 2113 | hCoV-19/England/MILK-A47DB1/2020\|EPI_ISL_598369\|20 | ···················· | ···················· | ····················· | ···················· | ······T············· | ···················· |
| N | 2114 | hCoV-19/England/MILK-A46AE8/2020\|EPI_ISL_598370\|20 | ···················· | ···················· | ····················· | ···················· | ···················· | ···················· |
| N | 2115 | hCoV-19/England/MILK-A47030/2020\|EPI_ISL_598371\|20 | ···················· | ···················· | ····················· | ···················· | ···················· | ···················· |
| N | 2116 | hCoV-19/England/MILK-A47E90/2020\|EPI_ISL_598373\|20 | ···················· | ···················· | ····················· | ···················· | ···················· | ···················· |
| N | 2117 | hCoV-19/England/MILK-A475B9/2020\|EPI_ISL_598374\|20 | ···················· | ···················· | ····················· | ···················· | ······T············· | ···················· |
| N | 2118 | hCoV-19/England/MILK-A4722B/2020\|EPI_ISL_598375\|20 | ···················· | ···················· | ····················· | ···················· | ···················· | ···················· |
| N | 2119 | hCoV-19/England/MILK-A47883/2020\|EPI_ISL_598376\|20 | ···················· | ····A··············· | ····················· | ···················· | ···················· | ···················· |
| N | 2120 | hCoV-19/England/MILK-A47355/2020\|EPI_ISL_598377\|20 | ···················· | ···················· | ····················· | ···················· | ···················· | ···················· |
| N | 2121 | hCoV-19/England/MILK-A4707C/2020\|EPI_ISL_598379\|20 | ···················· | ···················· | ····················· | ···················· | ···················· | ···················· |
| N | 2122 | hCoV-19/England/MILK-A47FBB/2020\|EPI_ISL_598380\|20 | ···················· | ···················· | ····················· | ···················· | ···················· | ···················· |
| N | 2123 | hCoV-19/England/MILK-A48011/2020\|EPI_ISL_598381\|20 | ···················· | ···················· | ····················· | ···················· | ···················· | ···················· |
| N | 2124 | hCoV-19/England/MILK-A47A05/2020\|EPI_ISL_598382\|20 | ···················· | ···················· | ····················· | ···················· | ···················· | ···················· |
| N | 2125 | hCoV-19/England/MILK-A46B21/2020\|EPI_ISL_598383\|20 | ···················· | ···················· | ····················· | ···················· | ···················· | ···················· |
| N | 2126 | hCoV-19/England/MILK-A470F4/2020\|EPI_ISL_598385\|20 | ···················· | ···················· | ····················· | ···················· | ···················· | ···················· |
| N | 2127 | hCoV-19/England/MILK-A47BE4/2020\|EPI_ISL_598386\|20 | ···················· | ···················· | ····················· | ···················· | ···················· | ···················· |
| N | 2128 | hCoV-19/England/MILK-A46AD9/2020\|EPI_ISL_598387\|20 | ···················· | ···················· | ····················· | ···················· | ···················· | ···················· |
| N | 2129 | hCoV-19/England/MILK-A469DC/2020\|EPI_ISL_598390\|20 | ···················· | ···················· | ····················· | ···················· | ···················· | ···················· |
| N | 2130 | hCoV-19/England/MILK-A4711F/2020\|EPI_ISL_598392\|20 | ···················· | ···················· | ····················· | ···················· | ···················· | ···················· |
| N | 2131 | hCoV-19/England/MILK-A46DEF/2020\|EPI_ISL_598393\|20 | ···················· | ···················· | ····················· | ···················· | ···················· | ···················· |
| N | 2132 | hCoV-19/England/MILK-A46972/2020\|EPI_ISL_598394\|20 | ···················· | ···················· | ····················· | ···················· | ···················· | ···················· |
| N | 2133 | hCoV-19/England/MILK-A469EB/2020\|EPI_ISL_598395\|20 | ···················· | ···················· | ····················· | ···················· | ···················· | ···················· |
| N | 2134 | hCoV-19/England/MILK-A47346/2020\|EPI_ISL_598396\|20 | ···················· | ···················· | ····················· | ···················· | ···················· | ···················· |
| N | 2135 | hCoV-19/England/MILK-A46E91/2020\|EPI_ISL_598397\|20 | ···················· | ···················· | ····················· | ···················· | ······T············· | ···················· |
| N | 2136 | hCoV-19/England/MILK-A476E3/2020\|EPI_ISL_598398\|20 | ···················· | ···················· | ····················· | ···················· | ···················· | ···················· |
| N | 2137 | hCoV-19/England/MILK-A473A0/2020\|EPI_ISL_598399\|20 | ···················· | ···················· | ····················· | ···················· | ···················· | ···················· |
| N | 2138 | hCoV-19/England/MILK-A47ABA/2020\|EPI_ISL_598402\|20 | ···················· | ···················· | ····················· | ···················· | ···················· | ···················· |
| N | 2139 | hCoV-19/England/MILK-A46F34/2020\|EPI_ISL_598403\|20 | ···················· | ···················· | ····················· | ···················· | ···················· | ···················· |
| N | 2140 | hCoV-19/England/MILK-A47452/2020\|EPI_ISL_598404\|20 | ···················· | ···················· | ····················· | ···················· | ···················· | ···················· |
| N | 2141 | hCoV-19/England/MILK-A46D1C/2020\|EPI_ISL_598405\|20 | ···················· | ···················· | ····················· | ···················· | ···················· | ···················· |
| N | 2142 | hCoV-19/England/MILK-A46A33/2020\|EPI_ISL_598406\|20 | ···················· | ···G················ | ····················· | ···················· | ···················· | ···················· |
| N | 2143 | hCoV-19/England/MILK-A47786/2020\|EPI_ISL_598407\|20 | ···················· | ···················· | ····················· | ···················· | ······T············· | ···················· |
| N | 2144 | hCoV-19/England/MILK-A47249/2020\|EPI_ISL_598410\|20 | ···················· | ···················· | ····················· | ···················· | ······T············· | ···················· |
| N | 2145 | hCoV-19/England/MILK-A46D49/2020\|EPI_ISL_598411\|20 | ···················· | ···················· | ····················· | ···················· | ···················· | ···················· |
| N | 2146 | hCoV-19/England/MILK-A47D2A/2020\|EPI_ISL_598412\|20 | ···················· | ···················· | ····················· | ···················· | ···················· | ···················· |
| N | 2147 | hCoV-19/England/MILK-A46F25/2020\|EPI_ISL_598425\|20 | ···················· | ···················· | ····················· | ···················· | ···················· | ···················· |

| | | | | | | | |
|---|---|---|---|---|---|---|---|
| N | 2148 hCoV-19/England/MILK-A46D67/2020\|EPI_ISL_598426\|20 | ···················· | ···················· | ····················· | ···················· | ···················· | ···················· |
| N | 2149 hCoV-19/England/MILK-A47DA2/2020\|EPI_ISL_598428\|20 | ···················· | ···················· | ····················· | ···················· | ······T············· | ···················· |
| N | 2150 hCoV-19/England/MILK-A47F7F/2020\|EPI_ISL_598429\|20 | ···················· | ···················· | ····················· | ···················· | ···················· | ···················· |
| N | 2151 hCoV-19/England/MILK-A474DA/2020\|EPI_ISL_598430\|20 | ···················· | ···················· | ····················· | ···················· | ···················· | ···················· |
| N | 2152 hCoV-19/England/MILK-A47DC0/2020\|EPI_ISL_598431\|20 | ···················· | ·A·················· | ····················· | ···················· | ···················· | ···················· |
| N | 2153 hCoV-19/England/MILK-A47258/2020\|EPI_ISL_598432\|20 | ···················· | ···················· | ····················· | ···················· | ···················· | ···················· |
| N | 2154 hCoV-19/England/MILK-A46CE2/2020\|EPI_ISL_598434\|20 | ···················· | ···················· | ····················· | ···················· | ···················· | ···················· |
| N | 2155 hCoV-19/England/MILK-A47B3F/2020\|EPI_ISL_598437\|20 | ···················· | ···················· | ····················· | ···················· | ···················· | ···················· |
| N | 2156 hCoV-19/England/MILK-A4765C/2020\|EPI_ISL_598438\|20 | ···················· | ···················· | ····················· | ···················· | ···················· | ···················· |
| N | 2157 hCoV-19/England/MILK-A46FBC/2020\|EPI_ISL_598439\|20 | ···················· | ···················· | ····················· | ···················· | ···················· | ···················· |
| N | 2158 hCoV-19/England/MILK-A46CF1/2020\|EPI_ISL_598440\|20 | ···················· | ···················· | ····················· | ···················· | ···················· | ···················· |
| N | 2159 hCoV-19/England/MILK-A46EBF/2020\|EPI_ISL_598441\|20 | ···················· | ···················· | ····················· | ···················· | ······T············· | ···················· |
| N | 2160 hCoV-19/England/MILK-A4808A/2020\|EPI_ISL_598442\|20 | ···················· | ···················· | ····················· | ···················· | ···················· | ···················· |
| N | 2161 hCoV-19/England/MILK-A4706D/2020\|EPI_ISL_598443\|20 | ···················· | ···················· | ····················· | ···················· | ···················· | ···················· |
| N | 2162 hCoV-19/England/MILK-A46C88/2020\|EPI_ISL_598444\|20 | ···················· | ···················· | ····················· | ···················· | ···················· | ···················· |
| N | 2163 hCoV-19/England/MILK-A4704F/2020\|EPI_ISL_598445\|20 | ···················· | ···················· | ····················· | ···················· | ···················· | ···················· |
| N | 2164 hCoV-19/England/QEUH-9DE530/2020\|EPI_ISL_599733\|20 | ···················· | ···················· | ····················· | ···················· | ···················· | ···················· |
| N | 2165 hCoV-19/England/QEUH-9DD0D6/2020\|EPI_ISL_599734\|20 | ···················· | ···················· | ····················· | ···················· | ···················· | ···················· |
| N | 2166 hCoV-19/Scotland/QEUH-9DD179/2020\|EPI_ISL_599735\|2 | ···················· | ···················· | ····················· | ···················· | ···················· | ···················· |
| N | 2167 hCoV-19/England/QEUH-9DDD1B/2020\|EPI_ISL_599736\|20 | ···················· | ···················· | ····················· | ···················· | ···················· | ···················· |
| N | 2168 hCoV-19/England/QEUH-9DCF8F/2020\|EPI_ISL_599738\|20 | ···················· | ·A·················· | ····················· | ···················· | ···················· | ···A················ |
| N | 2169 hCoV-19/England/QEUH-9DD1D3/2020\|EPI_ISL_599740\|20 | ···················· | ···················· | ····················· | ···················· | ···················· | ···················· |
| N | 2170 hCoV-19/England/QEUH-9DE05D/2020\|EPI_ISL_599741\|20 | ···················· | ···················· | ····················· | ···················· | ····T··············· | ···················· |
| N | 2171 hCoV-19/Scotland/QEUH-9DD3DD/2020\|EPI_ISL_599742\|2 | ···················· | ···················· | ····················· | ···················· | ···················· | ···················· |
| N | 2172 hCoV-19/England/QEUH-9DD461/2020\|EPI_ISL_599743\|20 | ···················· | ···················· | ····················· | ···················· | ···················· | ···················· |
| N | 2173 hCoV-19/England/QEUH-9DCD94/2020\|EPI_ISL_599755\|20 | ···················· | ···················· | ····················· | ···················· | ···················· | ···················· |
| N | 2174 hCoV-19/England/QEUH-9DE503/2020\|EPI_ISL_599756\|20 | ···················· | ·A·················· | ····················· | ···················· | ···················· | ···A················ |
| N | 2175 hCoV-19/England/QEUH-9DE309/2020\|EPI_ISL_599757\|20 | ···················· | ···················· | ····················· | ···················· | ···················· | ···················· |
| N | 2176 hCoV-19/England/QEUH-9DE248/2020\|EPI_ISL_599758\|20 | ···················· | ···················· | ····················· | ···················· | ···················· | ···················· |
| N | 2177 hCoV-19/England/QEUH-9DDD39/2020\|EPI_ISL_599759\|20 | ···················· | ···················· | ····················· | ···················· | ···················· | ···················· |
| N | 2178 hCoV-19/England/QEUH-9DD689/2020\|EPI_ISL_599761\|20 | ···················· | ···················· | ····················· | ···················· | ···················· | ···················· |
| N | 2179 hCoV-19/England/QEUH-9DCFF8/2020\|EPI_ISL_599762\|20 | ···················· | ···················· | ····················· | ···················· | ···················· | ···················· |
| N | 2180 hCoV-19/England/QEUH-9DE284/2020\|EPI_ISL_599764\|20 | ···················· | ···················· | ····················· | ···················· | ···················· | ···················· |
| N | 2181 hCoV-19/England/QEUH-9DD777/2020\|EPI_ISL_599766\|20 | ···················· | ···················· | ····················· | ···················· | ···················· | ···················· |
| N | 2182 hCoV-19/England/QEUH-9DE47F/2020\|EPI_ISL_599767\|20 | ···················· | ···················· | ····················· | ···················· | ···················· | ···················· |
| N | 2183 hCoV-19/England/QEUH-9DD892/2020\|EPI_ISL_599768\|20 | ···················· | ···················· | ····················· | ···················· | ···················· | ···················· |
| N | 2184 hCoV-19/England/QEUH-9DD9EA/2020\|EPI_ISL_599769\|20 | ···················· | ···················· | ····················· | ···················· | ···················· | ···················· |
| N | 2185 hCoV-19/Scotland/QEUH-9DD59B/2020\|EPI_ISL_599770\|2 | ···················· | ···················· | ····················· | ···················· | ···················· | ···················· |
| N | 2186 hCoV-19/England/QEUH-9DE1D2/2020\|EPI_ISL_599771\|20 | ···················· | ···················· | ····················· | ···················· | ······T············· | ···················· |
| N | 2187 hCoV-19/England/QEUH-9DDEDC/2020\|EPI_ISL_599772\|20 | ···················· | ···················· | ····················· | ···················· | ···················· | ···················· |
| N | 2188 hCoV-19/England/QEUH-9DD768/2020\|EPI_ISL_599773\|20 | ···················· | ···················· | ····················· | ···················· | ···················· | ···················· |
| N | 2189 hCoV-19/England/QEUH-9DD0F4/2020\|EPI_ISL_599774\|20 | ···················· | ···················· | ····················· | ···················· | ···················· | ···················· |
| N | 2190 hCoV-19/England/QEUH-9DE0A8/2020\|EPI_ISL_599775\|20 | ···················· | ···················· | ····················· | ···················· | ···················· | ···················· |
| N | 2191 hCoV-19/Scotland/QEUH-9DD56E/2020\|EPI_ISL_599776\|2 | ···················· | ···················· | ····················· | ···················· | ···················· | ···················· |
| N | 2192 hCoV-19/England/QEUH-9B8280/2020\|EPI_ISL_602140\|20 | ···················· | ·A·················· | ····················· | ···················· | ···················· | ···A················ |
| N | 2193 hCoV-19/England/QEUH-9B89F4/2020\|EPI_ISL_602141\|20 | ···················· | ···················· | ····················· | ···················· | ···················· | ···················· |
| N | 2194 hCoV-19/England/QEUH-9B8EC8/2020\|EPI_ISL_602142\|20 | ···················· | ···················· | ····················· | ···················· | ···················· | ···················· |
| N | 2195 hCoV-19/England/QEUH-9B81FC/2020\|EPI_ISL_602143\|20 | ···················· | ···················· | ····················· | ···················· | ···················· | ···················· |
| N | 2196 hCoV-19/England/QEUH-9B8842/2020\|EPI_ISL_602144\|20 | ···················· | ···················· | ····················· | ···················· | ···················· | ···················· |
| N | 2197 hCoV-19/Wales/MILK-9A6908/2020\|EPI_ISL_602145\|2020 | ···················· | ···················· | ····················· | ···················· | ···················· | ···················· |
| N | 2198 hCoV-19/Wales/MILK-99A5F4/2020\|EPI_ISL_602147\|2020 | ···················· | ···················· | ····················· | ···················· | ···················· | ···················· |
| N | 2199 hCoV-19/England/MILK-99DCA1/2020\|EPI_ISL_602148\|20 | ···················· | ···················· | ····················· | ···················· | ···················· | ···················· |
| N | 2200 hCoV-19/Wales/MILK-99DD9F/2020\|EPI_ISL_602149\|2020 | ···················· | ···················· | ····················· | ···················· | ···················· | ···················· |
| N | 2201 hCoV-19/England/MILK-99DCED/2020\|EPI_ISL_602151\|20 | ···················· | ···················· | ····················· | ···················· | ···················· | ···················· |
| N | 2202 hCoV-19/Wales/MILK-9AB51E/2020\|EPI_ISL_602152\|2020 | ···················· | ···················· | ····················· | ···················· | ···················· | ···················· |
| N | 2203 hCoV-19/Wales/MILK-993458/2020\|EPI_ISL_602153\|2020 | ···················· | ···················· | ····················· | ···················· | ···················· | ···················· |
| N | 2204 hCoV-19/Wales/CAMC-990CBD/2020\|EPI_ISL_602154\|2020 | ···················· | ···················· | ····················· | ···················· | ···················· | ···················· |
| N | 2205 hCoV-19/Wales/QEUH-96BB27/2020\|EPI_ISL_602156\|2020 | ···················· | ···················· | ····················· | ···················· | ···················· | ···················· |
| N | 2206 hCoV-19/Wales/ALDP-944113/2020\|EPI_ISL_602158\|2020 | ···················· | ···················· | ····················· | ···················· | ···················· | ···················· |
| N | 2207 hCoV-19/Wales/ALDP-944140/2020\|EPI_ISL_602159\|2020 | ···················· | ···················· | ····················· | ···················· | ···················· | ···················· |
| N | 2208 hCoV-19/England/ALDP-8E0095/2020\|EPI_ISL_602161\|20 | ···················· | ···················· | ····················· | ···················· | ···················· | ···················· |
| N | 2209 hCoV-19/USA/NM-DOH-2020307740/2020\|EPI_ISL_602162\| | ···················· | ···················· | ····················· | ···················· | ···················· | ···················· |
| N | 2210 hCoV-19/USA/NM-DOH-2020307741/2020\|EPI_ISL_602163\| | ···················· | ···················· | ····················· | ···················· | ···················· | ···················· |
| N | 2211 hCoV-19/USA/NM-DOH-2020307744/2020\|EPI_ISL_602164\| | ···················· | ···················· | ····················· | ···················· | ···················· | ···················· |
| N | 2212 hCoV-19/USA/NM-DOH-2020307857/2020\|EPI_ISL_602165\| | ···················· | ···················· | ····················· | ···················· | ···················· | ···················· |
| N | 2213 hCoV-19/USA/NM-DOH-2020307874/2020\|EPI_ISL_602166\| | ···················· | ·A·················· | ····················· | ···················· | ···················· | ···················· |
| N | 2214 hCoV-19/USA/NM-DOH-2020307954/2020\|EPI_ISL_602167\| | ···················· | ···················· | ····················· | ···················· | ···················· | ···················· |
| N | 2215 hCoV-19/USA/NM-DOH-2020307972/2020\|EPI_ISL_602168\| | ···················· | ····A··············· | ····················· | ···················· | ···················· | ···················· |
| N | 2216 hCoV-19/USA/NM-DOH-2020308002/2020\|EPI_ISL_602169\| | ···················· | ···················· | ····················· | ···················· | ···················· | ···················· |
| N | 2217 hCoV-19/USA/NM-DOH-2020308088/2020\|EPI_ISL_602170\| | ···················· | ···················· | ····················· | ···················· | ···················· | ···················· |
| N | 2218 hCoV-19/USA/NM-DOH-2020308126/2020\|EPI_ISL_602171\| | ···················· | ·A·················· | ····················· | ···················· | ···················· | ···················· |
| N | 2219 hCoV-19/USA/NM-DOH-2020308147/2020\|EPI_ISL_602172\| | ···················· | ···················· | ····················· | ···················· | ···················· | ···················· |
| N | 2220 hCoV-19/USA/NM-DOH-2020308229/2020\|EPI_ISL_602173\| | ···················· | ···················· | ····················· | ···················· | ···················· | ···················· |
| N | 2221 hCoV-19/USA/NM-DOH-2020308244/2020\|EPI_ISL_602174\| | ···················· | ···················· | ····················· | ···················· | ···················· | ···················· |
| N | 2222 hCoV-19/USA/NM-DOH-2020308245/2020\|EPI_ISL_602175\| | ···················· | ···················· | ····················· | ···················· | ···················· | ···················· |
| N | 2223 hCoV-19/USA/NM-DOH-2020308246/2020\|EPI_ISL_602176\| | ···················· | ···················· | ····················· | ···················· | ···················· | ···················· |
| N | 2224 hCoV-19/USA/NM-DOH-2020308298/2020\|EPI_ISL_602177\| | ···················· | ···················· | ····················· | ···················· | ···················· | ···················· |
| N | 2225 hCoV-19/USA/NM-DOH-2020308339/2020\|EPI_ISL_602178\| | ···················· | ···················· | ····················· | ···················· | ···················· | ···················· |
| N | 2226 hCoV-19/USA/NM-DOH-2020308427/2020\|EPI_ISL_602179\| | ···················· | ·A·················· | ····················· | ···················· | ···················· | ···················· |
| N | 2227 hCoV-19/USA/NM-DOH-2020308468/2020\|EPI_ISL_602180\| | ···················· | ···················· | ····················· | ···················· | ···················· | ···················· |

| | | | | | | | | |
|---|---|---|---|---|---|---|---|---|
| N | 2228 | hCoV-19/USA/NM-DOH-2020308490/2020\|EPI_ISL_602181\| | ···················· | ···················· | ···················· | ···················· | ···················· | ···················· |
| N | 2229 | hCoV-19/USA/NM-DOH-2020308494/2020\|EPI_ISL_602182\| | ···················· | ···················· | ···················· | ···················· | ···················· | ···················· |
| N | 2230 | hCoV-19/USA/NM-DOH-2020308662/2020\|EPI_ISL_602183\| | ···················· | ·A·················· | ···················· | ···················· | ···················· | ···················· |
| N | 2231 | hCoV-19/USA/NM-DOH-2020308663/2020\|EPI_ISL_602184\| | ···················· | ·A·················· | ···················· | ···················· | ···················· | ···················· |
| N | 2232 | hCoV-19/USA/NM-DOH-2020308703/2020\|EPI_ISL_602185\| | ···················· | ···················· | ···················· | ···················· | ···················· | ···················· |
| N | 2233 | hCoV-19/USA/NM-DOH-2020308708/2020\|EPI_ISL_602186\| | ···················· | ···················· | ···················· | ···················· | ···················· | ···················· |
| N | 2234 | hCoV-19/USA/NM-DOH-2020308907/2020\|EPI_ISL_602187\| | ···················· | ···················· | ···················· | ···················· | ···················· | ···················· |
| N | 2235 | hCoV-19/USA/NM-DOH-2020308915/2020\|EPI_ISL_602188\| | ···················· | ···················· | ···················· | ···················· | ···················· | ···················· |
| N | 2236 | hCoV-19/USA/NM-DOH-2020308923/2020\|EPI_ISL_602189\| | ···················· | ·A·················· | ···················· | ···················· | ···················· | ···················· |
| N | 2237 | hCoV-19/USA/NM-DOH-2020309011/2020\|EPI_ISL_602191\| | ···················· | ·A·················· | ···················· | ···················· | ···················· | ···················· |
| N | 2238 | hCoV-19/USA/NM-DOH-2020309015/2020\|EPI_ISL_602192\| | ···················· | ·A·················· | ···················· | ···················· | ···················· | ···················· |
| N | 2239 | hCoV-19/USA/NM-DOH-2020309108/2020\|EPI_ISL_602193\| | ···················· | ···················· | ···················· | ···················· | ···················· | ···················· |
| N | 2240 | hCoV-19/USA/NM-DOH-2020309699/2020\|EPI_ISL_602194\| | ···················· | ···················· | ···················· | ···················· | ···················· | ···················· |
| N | 2241 | hCoV-19/USA/NM-DOH-2020309716/2020\|EPI_ISL_602195\| | ···················· | ···················· | ···················· | ···················· | ···················· | ···················· |
| N | 2242 | hCoV-19/USA/NM-DOH-2020309719/2020\|EPI_ISL_602196\| | ···················· | ···················· | ···················· | ···················· | ···················· | ···················· |
| N | 2243 | hCoV-19/USA/NM-DOH-2020309735/2020\|EPI_ISL_602197\| | ···················· | ···················· | ···················· | ···················· | ···················· | ···················· |
| N | 2244 | hCoV-19/USA/NM-DOH-2020309744/2020\|EPI_ISL_602199\| | ···················· | ···················· | ···················· | ···················· | ···················· | ···················· |
| N | 2245 | hCoV-19/Switzerland/BE-ETHZ-300460/2020\|EPI_ISL_60 | ···················· | ···················· | ···················· | ···················· | ···················· | ···················· |
| N | 2246 | hCoV-19/Switzerland/BE-ETHZ-300462/2020\|EPI_ISL_60 | ···················· | ···················· | ···················· | ···················· | ···················· | ···················· |
| N | 2247 | hCoV-19/Switzerland/BE-ETHZ-300463/2020\|EPI_ISL_60 | ···················· | ···················· | ···················· | ···················· | ···················· | ···················· |
| N | 2248 | hCoV-19/Switzerland/VD-ETHZ-300465/2020\|EPI_ISL_60 | ···················· | ···················· | ···················· | ···················· | ···················· | ···················· |
| N | 2249 | hCoV-19/Switzerland/VD-ETHZ-300471/2020\|EPI_ISL_60 | ···················· | ···················· | ···················· | ···················· | ···················· | ···················· |
| N | 2250 | hCoV-19/Switzerland/UR-ETHZ-300472/2020\|EPI_ISL_60 | ···················· | ···················· | ···················· | ···················· | ···················· | ···················· |
| N | 2251 | hCoV-19/Switzerland/BS-ETHZ-300474/2020\|EPI_ISL_60 | ···················· | ···················· | ···················· | ···················· | ···················· | ···················· |
| N | 2252 | hCoV-19/Switzerland/BS-ETHZ-300475/2020\|EPI_ISL_60 | ···················· | ···················· | ···················· | ···················· | ···················· | ···················· |
| N | 2253 | hCoV-19/Switzerland/VD-ETHZ-300477/2020\|EPI_ISL_60 | ···················· | ···················· | ···················· | ···················· | ···················· | ···················· |
| N | 2254 | hCoV-19/Switzerland/ZH-ETHZ-310344/2020\|EPI_ISL_60 | ···················· | ···················· | ···················· | ···················· | ···················· | ···················· |
| N | 2255 | hCoV-19/Switzerland/ZH-ETHZ-310345/2020\|EPI_ISL_60 | ···················· | ···················· | ···················· | ···················· | ······K············· | ···················· |
| N | 2256 | hCoV-19/Switzerland/ZH-ETHZ-310346/2020\|EPI_ISL_60 | ···················· | ···················· | ···················· | ···················· | ···················· | ···················· |
| N | 2257 | hCoV-19/Switzerland/VD-ETHZ-310352/2020\|EPI_ISL_60 | ···················· | ···················· | ···················· | ···················· | ···················· | ···················· |
| N | 2258 | hCoV-19/Switzerland/VD-ETHZ-310353/2020\|EPI_ISL_60 | ···················· | ···················· | ···················· | ···················· | ···················· | ···················· |
| N | 2259 | hCoV-19/Switzerland/VD-ETHZ-310354/2020\|EPI_ISL_60 | ···················· | ···················· | ···················· | ···················· | ···················· | ···················· |
| N | 2260 | hCoV-19/Switzerland/AG-ETHZ-310396/2020\|EPI_ISL_60 | ···················· | ···················· | ···················· | ···················· | ···················· | ···················· |
| N | 2261 | hCoV-19/Switzerland/LU-ETHZ-310397/2020\|EPI_ISL_60 | ···················· | ···················· | ···················· | ···················· | ···················· | ···················· |
| N | 2262 | hCoV-19/Switzerland/SZ-ETHZ-310398/2020\|EPI_ISL_60 | ···················· | ···················· | ···················· | ···················· | ···················· | ···················· |
| N | 2263 | hCoV-19/Switzerland/SZ-ETHZ-310399/2020\|EPI_ISL_60 | ···················· | ···················· | ···················· | ···················· | ···················· | ···················· |
| N | 2264 | hCoV-19/Switzerland/SZ-ETHZ-310400/2020\|EPI_ISL_60 | ···················· | ···················· | ···················· | ···················· | ···················· | ···················· |
| N | 2265 | hCoV-19/Switzerland/AG-ETHZ-310401/2020\|EPI_ISL_60 | ···················· | ···················· | ···················· | ···················· | ···················· | ···················· |
| N | 2266 | hCoV-19/Switzerland/SZ-ETHZ-310402/2020\|EPI_ISL_60 | ···················· | ···················· | ···················· | ···················· | ···················· | ···················· |
| N | 2267 | hCoV-19/Switzerland/SZ-ETHZ-310403/2020\|EPI_ISL_60 | ···················· | ···················· | ···················· | ···················· | ···················· | ···················· |
| N | 2268 | hCoV-19/Switzerland/AG-ETHZ-310404/2020\|EPI_ISL_60 | ···················· | ···················· | ···················· | ···················· | ······K············· | ···················· |
| N | 2269 | hCoV-19/Switzerland/GR-ETHZ-310405/2020\|EPI_ISL_60 | ···················· | ···················· | ···················· | ···················· | ···················· | ···················· |
| N | 2270 | hCoV-19/Switzerland/GR-ETHZ-310406/2020\|EPI_ISL_60 | ···················· | ···················· | ···················· | ···················· | ······K············· | ···················· |
| N | 2271 | hCoV-19/Switzerland/AG-ETHZ-310407/2020\|EPI_ISL_60 | ···················· | ···················· | ···················· | ···················· | ···················· | ···················· |
| N | 2272 | hCoV-19/Switzerland/GR-ETHZ-310408/2020\|EPI_ISL_60 | ···················· | ···················· | ···················· | ···················· | ···················· | ···················· |
| N | 2273 | hCoV-19/Switzerland/SZ-ETHZ-310409/2020\|EPI_ISL_60 | ···················· | ···················· | ···················· | ···················· | ···················· | ···················· |
| N | 2274 | hCoV-19/Switzerland/SZ-ETHZ-310410/2020\|EPI_ISL_60 | ···················· | ···················· | ···················· | A··················· | ···················· | ···················· |
| N | 2275 | hCoV-19/Switzerland/GR-ETHZ-310411/2020\|EPI_ISL_60 | ···················· | ···················· | ···················· | ···················· | ···················· | ···················· |
| N | 2276 | hCoV-19/Switzerland/SZ-ETHZ-310412/2020\|EPI_ISL_60 | ···················· | ···················· | ···················· | ···················· | ···················· | ···················· |
| N | 2277 | hCoV-19/Switzerland/GR-ETHZ-310414/2020\|EPI_ISL_60 | ···················· | ···················· | ···················· | ···················· | ······K············· | ···················· |
| N | 2278 | hCoV-19/Switzerland/ZH-ETHZ-310415/2020\|EPI_ISL_60 | ···················· | ···················· | ···················· | ···················· | ······K············· | ···················· |
| N | 2279 | hCoV-19/Switzerland/ZH-ETHZ-310416/2020\|EPI_ISL_60 | ···················· | ···················· | ···················· | ···················· | ···················· | ···················· |
| N | 2280 | hCoV-19/Switzerland/ZH-ETHZ-310417/2020\|EPI_ISL_60 | ···················· | ···················· | ···················· | ···················· | ······K············· | ···················· |
| N | 2281 | hCoV-19/Switzerland/ZH-ETHZ-310418/2020\|EPI_ISL_60 | ···················· | ···················· | ···················· | ···················· | ···················· | ···················· |
| N | 2282 | hCoV-19/Switzerland/ZH-ETHZ-310419/2020\|EPI_ISL_60 | ···················· | ···················· | ···················· | ···················· | ···················· | ···················· |
| N | 2283 | hCoV-19/Switzerland/ZH-ETHZ-310420/2020\|EPI_ISL_60 | ···················· | ···················· | ···················· | ···················· | ······K············· | ···················· |
| N | 2284 | hCoV-19/Switzerland/ZH-ETHZ-310421/2020\|EPI_ISL_60 | ···················· | ···················· | ···················· | ···················· | ···················· | ···················· |
| N | 2285 | hCoV-19/Switzerland/ZH-ETHZ-310422/2020\|EPI_ISL_60 | ···················· | ···················· | ···················· | ···················· | ···················· | ···················· |
| N | 2286 | hCoV-19/Switzerland/ZH-ETHZ-310423/2020\|EPI_ISL_60 | ···················· | ···················· | ···················· | ···················· | ···················· | ···················· |
| N | 2287 | hCoV-19/Switzerland/ZH-ETHZ-310424/2020\|EPI_ISL_60 | ···················· | ···················· | ···················· | ···················· | ···················· | ···················· |
| N | 2288 | hCoV-19/Switzerland/ZH-ETHZ-310425/2020\|EPI_ISL_60 | ···················· | ···················· | ···················· | ···················· | ···················· | ···················· |
| N | 2289 | hCoV-19/USA/AZ-QDX-2448/2020\|EPI_ISL_604740\|2020-0 | ···················· | ···················· | ···················· | ···················· | ···················· | ···················· |
| N | 2290 | hCoV-19/USA/AZ-QDX-2525/2020\|EPI_ISL_604741\|2020-0 | ···················· | ···················· | ···················· | ···················· | ···················· | ···················· |
| N | 2291 | hCoV-19/USA/AZ-QDX-2547/2020\|EPI_ISL_604742\|2020-0 | ···················· | ···················· | ···················· | ···················· | ···················· | ···················· |
| N | 2292 | hCoV-19/USA/CA-QDX-2429/2020\|EPI_ISL_604743\|2020-0 | ···················· | ···················· | ···················· | ···················· | ···················· | ···················· |
| N | 2293 | hCoV-19/USA/CA-QDX-2433/2020\|EPI_ISL_604744\|2020-0 | ···················· | ···················· | ···················· | ···················· | ···················· | ···················· |
| N | 2294 | hCoV-19/USA/CA-QDX-2462/2020\|EPI_ISL_604749\|2020-0 | ···················· | ···················· | ···················· | ···················· | ···················· | ···················· |
| N | 2295 | hCoV-19/USA/CA-QDX-2470/2020\|EPI_ISL_604750\|2020-0 | ···················· | ···················· | ···················· | ···················· | ···················· | ···················· |
| N | 2296 | hCoV-19/USA/CA-QDX-2475/2020\|EPI_ISL_604751\|2020-0 | ···················· | ···················· | ···················· | ···················· | ···················· | ···················· |
| N | 2297 | hCoV-19/USA/CA-QDX-2477/2020\|EPI_ISL_604752\|2020-0 | ···················· | ···················· | ···················· | ···················· | ···················· | ···················· |
| N | 2298 | hCoV-19/USA/CA-QDX-2478/2020\|EPI_ISL_604753\|2020-0 | ···················· | ···················· | ···················· | ···················· | ···················· | ···················· |
| N | 2299 | hCoV-19/USA/CA-QDX-2485/2020\|EPI_ISL_604754\|2020-0 | ···················· | ···················· | ···················· | ···················· | ···················· | ···················· |
| N | 2300 | hCoV-19/USA/CA-QDX-2488/2020\|EPI_ISL_604755\|2020-0 | ···················· | ···················· | ···················· | ···················· | ···················· | ···················· |
| N | 2301 | hCoV-19/USA/CA-QDX-2489/2020\|EPI_ISL_604756\|2020-0 | ···················· | ···················· | ···················· | ···················· | ···················· | ···················· |
| N | 2302 | hCoV-19/USA/CA-QDX-2493/2020\|EPI_ISL_604757\|2020-0 | ···················· | ···················· | ···················· | ···················· | ···················· | ···················· |
| N | 2303 | hCoV-19/USA/CA-QDX-2496/2020\|EPI_ISL_604758\|2020-0 | ···················· | ···················· | ···················· | ···················· | ···················· | ···················· |
| N | 2304 | hCoV-19/USA/CA-QDX-2500/2020\|EPI_ISL_604759\|2020-0 | ···················· | ···················· | ···················· | ···················· | ···················· | ···················· |
| N | 2305 | hCoV-19/England/CAMC-A58685/2020\|EPI_ISL_597116\|20 | ···················· | ···················· | ···················· | ···················· | ···················· | ···················· |
| N | 2306 | hCoV-19/England/CAMC-A58FA8/2020\|EPI_ISL_597117\|20 | ···················· | ···················· | ···················· | ···················· | ···················· | ···················· |
| N | 2307 | hCoV-19/England/CAMC-A5903B/2020\|EPI_ISL_597118\|20 | ···················· | ···················· | ···················· | ···················· | ···················· | ···················· |

N 2308 hCoV-19/England/CAMC-A58A2F/2020|EPI_ISL_597119|20 ···················· ···················· ···················· ···················· ····T··············· ····················
N 2309 hCoV-19/England/CAMC-A58737/2020|EPI_ISL_597120|20 ···················· ···················· ···················· ···················· ···················· ····················
N 2310 hCoV-19/England/CAMC-A5853D/2020|EPI_ISL_597121|20 ···················· ···················· ···················· ···················· ······T············· ····················
N 2311 hCoV-19/England/CAMC-A5860D/2020|EPI_ISL_597122|20 ···················· ···················· ···················· ···················· ···················· ····················
N 2312 hCoV-19/England/CAMC-A586D0/2020|EPI_ISL_597124|20 ···················· ···················· ···················· ···················· ···················· ····················
N 2313 hCoV-19/England/CAMC-A59323/2020|EPI_ISL_597126|20 ···················· ···················· ···················· ···················· ···················· ····················
N 2314 hCoV-19/England/CAMC-A592EA/2020|EPI_ISL_597127|20 ···················· ···················· ···················· ···················· ···················· ····················
N 2315 hCoV-19/England/CAMC-A57F8B/2020|EPI_ISL_597128|20 ···················· ···················· ···················· ···················· ···················· ····················
N 2316 hCoV-19/England/CAMC-A59754/2020|EPI_ISL_597129|20 ···················· ···················· ···················· ···················· ···················· ····················
N 2317 hCoV-19/England/CAMC-A5810C/2020|EPI_ISL_597130|20 ···················· ···················· ···················· ···················· ···················· ····················
N 2318 hCoV-19/England/CAMC-A5838E/2020|EPI_ISL_597131|20 ···················· ···················· ···················· ···················· ···················· ····················
N 2319 hCoV-19/England/CAMC-A591DE/2020|EPI_ISL_597132|20 ···················· ···················· ···················· ···················· ···················· ····················
N 2320 hCoV-19/England/CAMC-A58B86/2020|EPI_ISL_597133|20 ···················· ···················· ···················· ···················· ···················· ····················
N 2321 hCoV-19/England/CAMC-A584F4/2020|EPI_ISL_597135|20 ···················· ···················· ···················· ···················· ···················· ····················
N 2322 hCoV-19/England/CAMC-A5861C/2020|EPI_ISL_597136|20 ···················· ···················· ···················· ···················· ······T············· ····················
N 2323 hCoV-19/England/CAMC-A5800F/2020|EPI_ISL_597137|20 ···················· ···················· ···················· ···················· ···················· ····················
N 2324 hCoV-19/England/CAMC-A59059/2020|EPI_ISL_597138|20 ···················· ···················· ···················· ···················· ···················· ····················
N 2325 hCoV-19/England/CAMC-A5988E/2020|EPI_ISL_597139|20 ···················· ···················· ···················· ···················· ···················· ····················
N 2326 hCoV-19/England/CAMC-A595C3/2020|EPI_ISL_597141|20 ···················· ···················· ···················· ···················· ···················· ····················
N 2327 hCoV-19/England/CAMC-A57D81/2020|EPI_ISL_597143|20 ···················· ···················· ···················· ···················· ···················· ····················
N 2328 hCoV-19/England/CAMC-A5895F/2020|EPI_ISL_597144|20 ···················· ···················· ···················· ···················· ···················· ····················
N 2329 hCoV-19/England/CAMC-A594C6/2020|EPI_ISL_597146|20 ···················· ·A·················· ···················· ···················· ···················· ···A················
N 2330 hCoV-19/England/CAMC-A58333/2020|EPI_ISL_597147|20 ···················· ···················· ···················· ···················· ···················· ····················
N 2331 hCoV-19/England/CAMC-A59499/2020|EPI_ISL_597148|20 ···················· ···················· ···················· ···················· ···················· ····················
N 2332 hCoV-19/England/CAMC-A5901D/2020|EPI_ISL_597149|20 ···················· ···················· ···················· ···················· ···················· ····················
N 2333 hCoV-19/England/CAMC-A5888F/2020|EPI_ISL_597150|20 ···················· ···················· ···················· ···················· ···················· ····················
N 2334 hCoV-19/England/CAMC-A57D90/2020|EPI_ISL_597151|20 ···················· ···················· ···················· ···················· ···················· ····················
N 2335 hCoV-19/England/CAMC-A58C83/2020|EPI_ISL_597152|20 ···················· ···················· ···················· ···················· ···················· ····················
N 2336 hCoV-19/England/CAMC-A58C1A/2020|EPI_ISL_597153|20 ···················· ···················· ···················· ···················· ···················· ····················
N 2337 hCoV-19/England/CAMC-A593D8/2020|EPI_ISL_597154|20 ···················· ···················· ···················· ···················· ···················· ····················
N 2338 hCoV-19/England/CAMC-A59068/2020|EPI_ISL_597155|20 ···················· ···················· ···················· ···················· ···················· ····················
N 2339 hCoV-19/England/CAMC-A58C47/2020|EPI_ISL_597156|20 ···················· ···················· ···················· ···················· ···················· ····················
N 2340 hCoV-19/England/CAMC-A5898C/2020|EPI_ISL_597157|20 ···················· ···················· ···················· ···················· ···················· ····················
N 2341 hCoV-19/England/MILK-A46A15/2020|EPI_ISL_598400|20 ···················· ···················· ···················· ···················· ···················· ····················
N 2342 hCoV-19/England/MILK-A474BC/2020|EPI_ISL_598401|20 ···················· ·A·················· ···················· ···················· ···················· ···A················
N 2343 hCoV-19/England/MILK-A47531/2020|EPI_ISL_598409|20 ···················· ···················· ···················· ···················· ···················· ····················
N 2344 hCoV-19/England/MILK-A46D58/2020|EPI_ISL_598414|20 ···················· ···················· ···················· ···················· ···················· ····················
N 2345 hCoV-19/England/MILK-A46CB5/2020|EPI_ISL_598415|20 ···················· ···················· ···················· ···················· ···················· ····················
N 2346 hCoV-19/England/MILK-A46C3D/2020|EPI_ISL_598416|20 ···················· ···················· ···················· ···················· ···················· ····················
N 2347 hCoV-19/England/MILK-A469AF/2020|EPI_ISL_598417|20 ···················· ···················· ···················· ···················· ···················· ····················
N 2348 hCoV-19/England/MILK-A47BB7/2020|EPI_ISL_598418|20 ···················· ···················· ···················· ···················· ···················· ····················
N 2349 hCoV-19/England/MILK-A46C97/2020|EPI_ISL_598419|20 ···················· ···················· ···················· ···················· ···················· ····················
N 2350 hCoV-19/England/MILK-A47364/2020|EPI_ISL_598420|20 ···················· ···················· ···················· ···················· ···················· ····················
N 2351 hCoV-19/England/MILK-A47EAF/2020|EPI_ISL_598421|20 ···················· ···················· ···················· ···················· ···················· ····················
N 2352 hCoV-19/England/MILK-A47D93/2020|EPI_ISL_598422|20 ···················· ···················· ···················· ···················· ···················· ····················
N 2353 hCoV-19/England/MILK-A46981/2020|EPI_ISL_598423|20 ···················· ···················· ···················· ···················· ···················· ····················
N 2354 hCoV-19/England/MILK-A469BE/2020|EPI_ISL_598446|20 ···················· ···················· ···················· ···················· ···················· ····················
N 2355 hCoV-19/England/MILK-A480C6/2020|EPI_ISL_598447|20 ···················· ···················· ···················· ···················· ···················· ····················
N 2356 hCoV-19/England/MILK-A47513/2020|EPI_ISL_598448|20 ···················· ·A·················· ···················· ···················· ···················· ····················
N 2357 hCoV-19/England/MILK-A47AE7/2020|EPI_ISL_598449|20 ···················· ···················· ···················· ···················· ···················· ····················
N 2358 hCoV-19/England/MILK-A48169/2020|EPI_ISL_598450|20 ···················· ···················· ···················· ···················· ···················· ····················
N 2359 hCoV-19/England/MILK-A47D39/2020|EPI_ISL_598451|20 ···················· ···················· ···················· ···················· ···················· ····················
N 2360 hCoV-19/England/MILK-A475D7/2020|EPI_ISL_598453|20 ···················· ···················· ···················· ···················· ···················· ····················
N 2361 hCoV-19/England/MILK-A469CD/2020|EPI_ISL_598454|20 ···················· ···················· ···················· ···················· ···················· ····················
N 2362 hCoV-19/England/MILK-A4763E/2020|EPI_ISL_598455|20 ···················· ···················· ···················· ···················· ···················· ····················
N 2363 hCoV-19/England/MILK-A46936/2020|EPI_ISL_598456|20 ···················· ···················· ···················· ···················· ······T············· ···············A····
N 2364 hCoV-19/England/MILK-A46D3A/2020|EPI_ISL_598457|20 ···················· ···················· ···················· ···················· ···················· ····················
N 2365 hCoV-19/England/MILK-A46E37/2020|EPI_ISL_598459|20 ···················· ···················· ···················· ···················· ···················· ····················
N 2366 hCoV-19/England/MILK-A46954/2020|EPI_ISL_598460|20 ···················· ···················· ···················· ···················· ······T············· ····················
N 2367 hCoV-19/England/MILK-A46B6D/2020|EPI_ISL_598461|20 ···················· ···················· ···················· ···················· ···················· ····················
N 2368 hCoV-19/England/MILK-A471B5/2020|EPI_ISL_598463|20 ···················· ···················· ···················· ···················· ···················· ····················
N 2369 hCoV-19/England/MILK-A46927/2020|EPI_ISL_598464|20 ···················· ···················· ···················· ···················· ···················· ····················
N 2370 hCoV-19/England/MILK-A47A23/2020|EPI_ISL_598465|20 ···················· ···················· ···················· ···················· ···················· ····················
N 2371 hCoV-19/England/MILK-A46FF8/2020|EPI_ISL_598466|20 ···················· ···················· ···················· ···················· ······T············· ····················
N 2372 hCoV-19/England/MILK-A47F9D/2020|EPI_ISL_598467|20 ···················· ···················· ···················· ···················· ···················· ····················
N 2373 hCoV-19/England/MILK-A47962/2020|EPI_ISL_598468|20 ···················· ···················· ···················· ···················· ······T············· ····················
N 2374 hCoV-19/England/MILK-A472C1/2020|EPI_ISL_598469|20 ···················· ···················· ···················· ···················· ···················· ····················
N 2375 hCoV-19/England/MILK-A47E18/2020|EPI_ISL_598470|20 ···················· ···················· ···················· ···················· ······T············· ····················
N 2376 hCoV-19/England/MILK-A47425/2020|EPI_ISL_598471|20 ···················· ···················· ···················· ···················· ···················· ····················
N 2377 hCoV-19/England/MILK-A47B6C/2020|EPI_ISL_598472|20 ···················· ···················· ···················· ···················· ···················· ····················
N 2378 hCoV-19/England/MILK-A46ABB/2020|EPI_ISL_598473|20 ···················· ···················· ···················· ···················· ···················· ····················
N 2379 hCoV-19/England/MILK-A47197/2020|EPI_ISL_598474|20 ···················· ···················· ···················· ···················· ···················· ····················
N 2380 hCoV-19/England/MILK-A475C8/2020|EPI_ISL_598475|20 ···················· ···················· ···················· ···················· ···················· ····················
N 2381 hCoV-19/England/MILK-A47BC6/2020|EPI_ISL_598476|20 ···················· ···················· ···················· ···················· ···················· ····················
N 2382 hCoV-19/England/MILK-A47F06/2020|EPI_ISL_598478|20 ···················· ···················· ···················· ···················· ······T············· ····················
N 2383 hCoV-19/England/MILK-A47100/2020|EPI_ISL_598479|20 ···················· ···················· ···················· ···················· ···················· ····················
N 2384 hCoV-19/England/MILK-A47391/2020|EPI_ISL_598481|20 ···················· ···················· ···················· ···················· ···················· ····················
N 2385 hCoV-19/England/MILK-A47B11/2020|EPI_ISL_598482|20 ···················· ···················· ···················· ···················· ···················· ····················
N 2386 hCoV-19/England/MILK-A472FE/2020|EPI_ISL_598483|20 ···················· ···················· ···················· ···················· ···················· ····················
N 2387 hCoV-19/England/MILK-A47337/2020|EPI_ISL_598484|20 ···················· ···················· ···················· ···················· ···················· ····················

| | | | | | | | | |
|---|---|---|---|---|---|---|---|---|
| N | 2388 | hCoV-19/England/MILK-A47601/2020|EPI_ISL_598485|20 | ···················· | ·A·················· | ···················· | ···················· | ···················· | ···A················ |
| N | 2389 | hCoV-19/England/MILK-A480B7/2020|EPI_ISL_598486|20 | ···················· | ···················· | ···················· | ···················· | ···················· | ···················· |
| N | 2390 | hCoV-19/England/MILK-A478DE/2020|EPI_ISL_598487|20 | ···················· | ···················· | ···················· | ···················· | ···················· | ···················· |
| N | 2391 | hCoV-19/England/MILK-A47874/2020|EPI_ISL_598488|20 | ···················· | ···················· | ···················· | ···················· | ······T············· | ···················· |
| N | 2392 | hCoV-19/Scotland/QEUH-9DD188/2020|EPI_ISL_599701|2 | ···················· | ···················· | ···················· | ···················· | ···················· | ···················· |
| N | 2393 | hCoV-19/Scotland/QEUH-9DD382/2020|EPI_ISL_599702|2 | ···················· | ···················· | ···················· | ···················· | ···················· | ···················· |
| N | 2394 | hCoV-19/England/QEUH-9DDB3F/2020|EPI_ISL_599703|20 | ···················· | ···················· | ···················· | ···················· | ···················· | ···················· |
| N | 2395 | hCoV-19/Scotland/QEUH-9DD55F/2020|EPI_ISL_599704|2 | ···················· | ···················· | ···················· | ···················· | ···················· | ···················· |
| N | 2396 | hCoV-19/England/QEUH-9DD8DE/2020|EPI_ISL_599705|20 | ···················· | ···················· | ···················· | ···················· | ···················· | ···················· |
| N | 2397 | hCoV-19/England/QEUH-9DE1C3/2020|EPI_ISL_599706|20 | ···················· | ···················· | ···················· | ···················· | ···················· | ···················· |
| N | 2398 | hCoV-19/England/QEUH-9DCF25/2020|EPI_ISL_599707|20 | ···················· | ···················· | ···················· | ···················· | ···················· | ···················· |
| N | 2399 | hCoV-19/England/QEUH-9DDA32/2020|EPI_ISL_599708|20 | ···················· | ···················· | ···················· | ···················· | ····T··············· | ···················· |
| N | 2400 | hCoV-19/England/QEUH-9DD63E/2020|EPI_ISL_599709|20 | ···················· | ···················· | ···················· | ···················· | ····T··············· | ···················· |
| N | 2401 | hCoV-19/England/QEUH-9DE13C/2020|EPI_ISL_599710|20 | ···················· | ···················· | ···················· | ···················· | ···················· | ···················· |
| N | 2402 | hCoV-19/England/QEUH-9DE3BE/2020|EPI_ISL_599711|20 | ···················· | ···················· | ···················· | ···················· | ···················· | ···················· |
| N | 2403 | hCoV-19/England/QEUH-9DE2C0/2020|EPI_ISL_599712|20 | ···················· | ···················· | ···················· | ···················· | ···················· | ···················· |
| N | 2404 | hCoV-19/Wales/QEUH-9DD8CF/2020|EPI_ISL_599713|2020 | ···················· | ·A·················· | ···················· | ···················· | ···················· | ···A················ |
| N | 2405 | hCoV-19/England/QEUH-9DD81A/2020|EPI_ISL_599714|20 | ···················· | ···················· | ···················· | ···················· | ···················· | ···················· |
| N | 2406 | hCoV-19/England/QEUH-9DDE81/2020|EPI_ISL_599715|20 | ···················· | ···················· | ···················· | ···················· | ····T··············· | ···················· |
| N | 2407 | hCoV-19/England/QEUH-9DD0C7/2020|EPI_ISL_599716|20 | ···················· | ···················· | ···················· | ···················· | ···················· | ···················· |
| N | 2408 | hCoV-19/England/QEUH-9DDB8A/2020|EPI_ISL_599717|20 | ···················· | ···················· | ···················· | ···················· | ···················· | ···················· |
| N | 2409 | hCoV-19/Scotland/QEUH-9DD3EC/2020|EPI_ISL_599718|2 | ···················· | ···················· | ···················· | ···················· | ···················· | ···················· |
| N | 2410 | hCoV-19/England/QEUH-9DD829/2020|EPI_ISL_599719|20 | ···················· | ···················· | ···················· | ···················· | ···················· | ···················· |
| N | 2411 | hCoV-19/England/QEUH-9DE415/2020|EPI_ISL_599720|20 | ···················· | ·A·················· | ···················· | ···················· | ···················· | ···A················ |
| N | 2412 | hCoV-19/Scotland/QEUH-9DD540/2020|EPI_ISL_599721|2 | ···················· | ···················· | ···················· | ···················· | ···················· | ···················· |
| N | 2413 | hCoV-19/Scotland/QEUH-9DD8B0/2020|EPI_ISL_599722|2 | ···················· | ···················· | ···················· | ···················· | ···················· | ···················· |
| N | 2414 | hCoV-19/Scotland/QEUH-9DD4E9/2020|EPI_ISL_599723|2 | ···················· | ···················· | ···················· | ···················· | ···················· | ···················· |
| N | 2415 | hCoV-19/England/QEUH-9DDD48/2020|EPI_ISL_599724|20 | ···················· | ·A·················· | ···················· | ···················· | ···················· | ···A················ |
| N | 2416 | hCoV-19/Scotland/QEUH-9DDB4E/2020|EPI_ISL_599725|2 | ···················· | ···················· | ···················· | ···················· | ···················· | ···················· |
| N | 2417 | hCoV-19/Scotland/QEUH-9DD4BC/2020|EPI_ISL_599726|2 | ···················· | ···················· | ···················· | ···················· | ···················· | ···················· |
| N | 2418 | hCoV-19/England/QEUH-9DE4BB/2020|EPI_ISL_599727|20 | ···················· | ···················· | ···················· | ···················· | ···················· | ···················· |
| N | 2419 | hCoV-19/England/QEUH-9DDB02/2020|EPI_ISL_599728|20 | ···················· | ···················· | ···················· | ···················· | ···················· | ···················· |
| N | 2420 | hCoV-19/England/QEUH-9DD5D7/2020|EPI_ISL_599729|20 | ···················· | ···················· | ···················· | ···················· | ···················· | ···················· |
| N | 2421 | hCoV-19/England/QEUH-9DCBE5/2020|EPI_ISL_599732|20 | ···················· | ···················· | ···················· | ···················· | ···················· | ···················· |
| N | 2422 | hCoV-19/Scotland/QEUH-9DD7A4/2020|EPI_ISL_599737|2 | ···················· | ···················· | ···················· | ···················· | ···················· | ···················· |
| N | 2423 | hCoV-19/England/QEUH-9DDEBE/2020|EPI_ISL_599739|20 | ···················· | ···················· | ···················· | ···················· | ······T············· | ···················· |
| N | 2424 | hCoV-19/England/QEUH-9DE460/2020|EPI_ISL_599744|20 | ···················· | ···················· | ···················· | ···················· | ···················· | ···················· |
| N | 2425 | hCoV-19/Scotland/QEUH-9DDBF3/2020|EPI_ISL_599745|2 | ···················· | ···················· | ···················· | ···················· | ···················· | ···················· |
| N | 2426 | hCoV-19/Scotland/QEUH-9DD4CB/2020|EPI_ISL_599746|2 | ···················· | ···················· | ···················· | ···················· | ···················· | ···················· |
| N | 2427 | hCoV-19/England/QEUH-9DCF61/2020|EPI_ISL_599747|20 | ···················· | ·A·················· | ···················· | ···················· | ···················· | ···A················ |
| N | 2428 | hCoV-19/England/QEUH-9DD926/2020|EPI_ISL_599748|20 | ···················· | ···················· | ···················· | ···················· | ···················· | ···················· |
| N | 2429 | hCoV-19/England/QEUH-9DDD57/2020|EPI_ISL_599749|20 | ···················· | ·A·················· | ···················· | ···················· | ···················· | ···A················ |
| N | 2430 | hCoV-19/England/QEUH-9DD06D/2020|EPI_ISL_599750|20 | ···················· | ···················· | ···················· | ···················· | ···················· | ···················· |
| N | 2431 | hCoV-19/England/QEUH-9DDBD5/2020|EPI_ISL_599751|20 | ···················· | ···················· | ··········T········· | ···················· | ···················· | ···················· |
| N | 2432 | hCoV-19/England/QEUH-9DE187/2020|EPI_ISL_599752|20 | ···················· | ···················· | ···················· | ···················· | ···················· | ···················· |
| N | 2433 | hCoV-19/England/QEUH-9DD759/2020|EPI_ISL_599753|20 | ···················· | ···················· | ···················· | ···················· | ···················· | ···················· |
| N | 2434 | hCoV-19/England/QEUH-9DD5AA/2020|EPI_ISL_599754|20 | ···················· | ···················· | ···················· | ···················· | ···················· | ···················· |
| N | 2435 | hCoV-19/England/QEUH-9DE21B/2020|EPI_ISL_599778|20 | ···················· | ···················· | ···················· | ···················· | ···················· | ···················· |
| N | 2436 | hCoV-19/Scotland/QEUH-9DDA05/2020|EPI_ISL_599779|2 | ···················· | ···················· | ···················· | ···················· | ···················· | ···················· |
| N | 2437 | hCoV-19/England/QEUH-9DD416/2020|EPI_ISL_599780|20 | ···················· | ···················· | ···················· | ···················· | ···················· | ···················· |
| N | 2438 | hCoV-19/England/QEUH-9DD197/2020|EPI_ISL_599781|20 | ···················· | ···················· | ···················· | ···················· | ···················· | ···················· |
| N | 2439 | hCoV-19/England/QEUH-9DE433/2020|EPI_ISL_599782|20 | ···················· | ·A·················· | ···················· | ···················· | ···················· | ···A················ |
| N | 2440 | hCoV-19/England/QEUH-9DE8CE/2020|EPI_ISL_599785|20 | ···················· | ·A·················· | ···················· | ···················· | ···················· | ···A················ |
| N | 2441 | hCoV-19/Scotland/QEUH-9DD294/2020|EPI_ISL_599786|2 | ···················· | ···················· | ···················· | ···················· | ···················· | ···················· |
| N | 2442 | hCoV-19/England/QEUH-9DE63D/2020|EPI_ISL_599787|20 | ···················· | ···················· | ···················· | ···················· | ···················· | ···················· |
| N | 2443 | hCoV-19/England/QEUH-9DDC3C/2020|EPI_ISL_599789|20 | ···················· | ···················· | ···················· | ···················· | ···················· | ···················· |
| N | 2444 | hCoV-19/England/QEUH-9DDDEE/2020|EPI_ISL_599790|20 | ···················· | ···················· | ···················· | ···················· | ···················· | ···················· |
| N | 2445 | hCoV-19/England/QEUH-9DE011/2020|EPI_ISL_599791|20 | ···················· | ···················· | ···················· | ···················· | ···················· | ···················· |
| N | 2446 | hCoV-19/England/QEUH-9DD0E5/2020|EPI_ISL_599792|20 | ···················· | ···················· | ···················· | ···················· | ···················· | ···················· |
| N | 2447 | hCoV-19/Scotland/QEUH-9DD57D/2020|EPI_ISL_599793|2 | ···················· | ···················· | ···················· | ···················· | ···················· | ···················· |
| N | 2448 | hCoV-19/England/QEUH-9DDA23/2020|EPI_ISL_599794|20 | ···················· | ···················· | ···················· | ···················· | ···················· | ···················· |
| N | 2449 | hCoV-19/England/QEUH-9DDDC0/2020|EPI_ISL_599795|20 | ···················· | ···················· | ···················· | ···················· | ···················· | ···················· |
| N | 2450 | hCoV-19/England/QEUH-9DD5E6/2020|EPI_ISL_599796|20 | ···················· | ···················· | ···················· | ···················· | ···················· | ···················· |
| N | 2451 | hCoV-19/England/QEUH-9DDF9D/2020|EPI_ISL_599797|20 | ···················· | ···················· | ···················· | ···················· | ···················· | ···················· |
| N | 2452 | hCoV-19/England/QEUH-9DDEEB/2020|EPI_ISL_599798|20 | ···················· | ···················· | ···················· | ···················· | ······T············· | ···················· |
| N | 2453 | hCoV-19/England/QEUH-9DD030/2020|EPI_ISL_599799|20 | ···················· | ···················· | ···················· | ···················· | ···················· | ···················· |
| N | 2454 | hCoV-19/Scotland/QEUH-9CEA61/2020|EPI_ISL_602001|2 | ···················· | ···················· | ···················· | ···················· | ···················· | ···················· |
| N | 2455 | hCoV-19/Scotland/QEUH-9CF480/2020|EPI_ISL_602002|2 | ···················· | ···················· | ···················· | ···················· | ···················· | ···················· |
| N | 2456 | hCoV-19/Scotland/QEUH-9CF7D2/2020|EPI_ISL_602004|2 | ···················· | ···················· | ···················· | ···················· | ···················· | ···················· |
| N | 2457 | hCoV-19/Scotland/QEUH-9CF3A1/2020|EPI_ISL_602005|2 | ···················· | ···················· | ···················· | ···················· | ···················· | ···················· |
| N | 2458 | hCoV-19/Scotland/QEUH-9CE7A6/2020|EPI_ISL_602007|2 | ···················· | ···················· | ···················· | ···················· | ···················· | ···················· |
| N | 2459 | hCoV-19/Scotland/QEUH-9CE3EE/2020|EPI_ISL_602008|2 | ···················· | ···················· | ···················· | ···················· | ···················· | ···················· |
| N | 2460 | hCoV-19/Scotland/QEUH-9CECC5/2020|EPI_ISL_602009|2 | ···················· | ···················· | ···················· | ···················· | ···················· | ···················· |
| N | 2461 | hCoV-19/Scotland/QEUH-9CEECF/2020|EPI_ISL_602012|2 | ···················· | ···················· | ···················· | ···················· | ···················· | ···················· |
| N | 2462 | hCoV-19/Scotland/QEUH-9CE3C0/2020|EPI_ISL_602013|2 | ···················· | ···················· | ···················· | ···················· | ···················· | ···················· |
| N | 2463 | hCoV-19/Scotland/QEUH-9CEFBD/2020|EPI_ISL_602014|2 | ···················· | ···················· | ···················· | ···················· | ···················· | ···················· |
| N | 2464 | hCoV-19/Scotland/QEUH-9CF82A/2020|EPI_ISL_602016|2 | ···················· | ···················· | ···················· | ···················· | ···················· | ···················· |
| N | 2465 | hCoV-19/Scotland/QEUH-9CEF26/2020|EPI_ISL_602017|2 | ···················· | ···················· | ···················· | ···················· | ···················· | ···················· |
| N | 2466 | hCoV-19/Scotland/QEUH-9CF769/2020|EPI_ISL_602018|2 | ···················· | ···················· | ···················· | ···················· | ···················· | ···················· |
| N | 2467 | hCoV-19/Scotland/QEUH-9CE9BF/2020|EPI_ISL_602019|2 | ···················· | ···················· | ···················· | ···················· | ···················· | ···················· |

| | | | | | | | | | |
|---|---|---|---|---|---|---|---|---|---|
| N | 2468 | hCoV-19/Switzerland/BE-ETHZ-300276/2020\|EPI_ISL_60 | ···················· | ···················· | ···················· | ···················· | ···················· | ···················· | |
| N | 2469 | hCoV-19/Switzerland/BE-ETHZ-300277/2020\|EPI_ISL_60 | ···················· | ···················· | ···················· | ···················· | ······K············· | ···················· | |
| N | 2470 | hCoV-19/Switzerland/BE-ETHZ-300278/2020\|EPI_ISL_60 | ···················· | ···················· | ···················· | ···················· | ···················· | ···················· | |
| N | 2471 | hCoV-19/Switzerland/BE-ETHZ-300279/2020\|EPI_ISL_60 | ···················· | ···················· | ···················· | ···················· | ···················· | ···················· | |
| N | 2472 | hCoV-19/Switzerland/BE-ETHZ-300280/2020\|EPI_ISL_60 | ···················· | ···················· | ···················· | ···················· | ···················· | ···················· | |
| N | 2473 | hCoV-19/Switzerland/BE-ETHZ-300281/2020\|EPI_ISL_60 | ···················· | ···················· | ···················· | ···················· | ···················· | ···················· | |
| N | 2474 | hCoV-19/Switzerland/GE-ETHZ-300282/2020\|EPI_ISL_60 | ···················· | ···················· | ···················· | ···················· | ···················· | ···················· | |
| N | 2475 | hCoV-19/Switzerland/ZH-ETHZ-300283/2020\|EPI_ISL_60 | ···················· | ···················· | ···················· | ···················· | ···················· | ···················· | |
| N | 2476 | hCoV-19/Switzerland/ZH-ETHZ-300284/2020\|EPI_ISL_60 | ···················· | ···················· | ···················· | ···················· | ···················· | ···················· | |
| N | 2477 | hCoV-19/Switzerland/FR-ETHZ-300286/2020\|EPI_ISL_60 | ···················· | ···················· | ···················· | ···················· | ···················· | ···················· | |
| N | 2478 | hCoV-19/Switzerland/BE-ETHZ-300287/2020\|EPI_ISL_60 | ···················· | ···················· | ···················· | ···················· | ···················· | ···················· | |
| N | 2479 | hCoV-19/Switzerland/BE-ETHZ-300288/2020\|EPI_ISL_60 | ···················· | ···················· | ···················· | ···················· | ···················· | ···················· | |
| N | 2480 | hCoV-19/Switzerland/BE-ETHZ-300289/2020\|EPI_ISL_60 | ···················· | ···················· | ···················· | ···················· | ···················· | ···················· | |
| N | 2481 | hCoV-19/Switzerland/SG-ETHZ-300290/2020\|EPI_ISL_60 | ···················· | ···················· | ···················· | ···················· | ···················· | ···················· | |
| N | 2482 | hCoV-19/Switzerland/BE-ETHZ-300291/2020\|EPI_ISL_60 | ···················· | ···················· | ···················· | ···················· | ···················· | ···················· | |
| N | 2483 | hCoV-19/Switzerland/GE-ETHZ-300292/2020\|EPI_ISL_60 | ···················· | ···················· | ···················· | ···················· | ···················· | ···················· | |
| N | 2484 | hCoV-19/Switzerland/GE-ETHZ-300293/2020\|EPI_ISL_60 | ···················· | ···················· | ···················· | ···················· | ···················· | ···················· | |
| N | 2485 | hCoV-19/Switzerland/VD-ETHZ-300295/2020\|EPI_ISL_60 | ···················· | ···················· | ···················· | ···················· | ···················· | ···················· | |
| N | 2486 | hCoV-19/Switzerland/BE-ETHZ-300296/2020\|EPI_ISL_60 | ···················· | ···················· | ···················· | ···················· | ···················· | ···················· | |
| N | 2487 | hCoV-19/Switzerland/VD-ETHZ-300298/2020\|EPI_ISL_60 | ···················· | ···················· | ···················· | ···················· | ···················· | ···················· | |
| N | 2488 | hCoV-19/Switzerland/ZH-ETHZ-300299/2020\|EPI_ISL_60 | ···················· | ···················· | ···················· | ···················· | ···················· | ···················· | |
| N | 2489 | hCoV-19/Switzerland/BE-ETHZ-300300/2020\|EPI_ISL_60 | ···················· | ···················· | ···················· | ···················· | ···················· | ···················· | |
| N | 2490 | hCoV-19/Switzerland/VD-ETHZ-300301/2020\|EPI_ISL_60 | ···················· | ···················· | ···················· | ···················· | ···················· | ···················· | |
| N | 2491 | hCoV-19/Switzerland/VD-ETHZ-300302/2020\|EPI_ISL_60 | ···················· | ···················· | ···················· | ···················· | ···················· | ···················· | |
| N | 2492 | hCoV-19/Switzerland/VD-ETHZ-300303/2020\|EPI_ISL_60 | ···················· | ···················· | ···················· | ···················· | ···················· | ···················· | |
| N | 2493 | hCoV-19/Switzerland/ZH-ETHZ-300304/2020\|EPI_ISL_60 | ···················A | ···················· | ···················· | ···················· | ···················· | ···················· | 1 |
| N | 2494 | hCoV-19/Switzerland/VD-ETHZ-300305/2020\|EPI_ISL_60 | ···················· | ···················· | ···················· | ···················· | ···················· | ···················· | |
| N | 2495 | hCoV-19/Switzerland/BE-ETHZ-300306/2020\|EPI_ISL_60 | ···················· | ···················· | ···················· | ···················· | ···················· | ···················· | |
| N | 2496 | hCoV-19/Switzerland/BE-ETHZ-300307/2020\|EPI_ISL_60 | ···················· | ···················· | ···················· | ···················· | ···················· | ···················· | |
| N | 2497 | hCoV-19/Switzerland/VD-ETHZ-300308/2020\|EPI_ISL_60 | ···················· | ···················· | ···················· | ···················· | ···················· | ···················· | |
| N | 2498 | hCoV-19/Switzerland/BL-ETHZ-300309/2020\|EPI_ISL_60 | ···················· | ···················· | ···················· | ···················· | ···················· | ···················· | |
| N | 2499 | hCoV-19/Switzerland/BL-ETHZ-300310/2020\|EPI_ISL_60 | ···················· | ···················· | ···················· | ···················· | ···················· | ···················· | |
| N | 2500 | hCoV-19/Switzerland/BS-ETHZ-300311/2020\|EPI_ISL_60 | ···················· | ···················· | ···················· | ···················· | ···················· | ···················· | |
| N | 2501 | hCoV-19/Switzerland/AG-ETHZ-300312/2020\|EPI_ISL_60 | ···················· | ···················· | ···················· | ···················· | ···················· | ···················· | |
| N | 2502 | hCoV-19/Switzerland/BL-ETHZ-300313/2020\|EPI_ISL_60 | ···················· | ···················· | ···················· | ···················· | ···················· | ···················· | |
| N | 2503 | hCoV-19/Switzerland/GE-ETHZ-300314/2020\|EPI_ISL_60 | ···················· | ···················· | ···················· | ···················· | ···················· | ···················· | |
| N | 2504 | hCoV-19/Switzerland/VD-ETHZ-300316/2020\|EPI_ISL_60 | ···················· | ···················· | ···················· | ···················· | ···················· | ···················· | |
| N | 2505 | hCoV-19/USA/CA-QDX-2337/2020\|EPI_ISL_604639\|2020-0 | ···················· | ···················· | ···················· | ···················· | ···················· | ···················· | |
| N | 2506 | hCoV-19/USA/CA-QDX-2338/2020\|EPI_ISL_604640\|2020-0 | ···················· | ···················· | ···················· | ···················· | ···················· | ···················· | |
| N | 2507 | hCoV-19/USA/CA-QDX-2340/2020\|EPI_ISL_604641\|2020-0 | ···················· | ···················· | ···················· | ···················· | ···················· | ···················· | |
| N | 2508 | hCoV-19/USA/CA-QDX-2341/2020\|EPI_ISL_604642\|2020-0 | ···················· | ···················· | ···················· | ···················· | ···················· | ···················· | |
| N | 2509 | hCoV-19/USA/CA-QDX-2342/2020\|EPI_ISL_604643\|2020-0 | ···················· | ···················· | ···················· | ···················· | ···················· | ···················· | |
| N | 2510 | hCoV-19/USA/CA-QDX-2343/2020\|EPI_ISL_604644\|2020-0 | ···················· | ···················· | ···················· | ···················· | ···················· | ···················· | |
| N | 2511 | hCoV-19/USA/CA-QDX-2344/2020\|EPI_ISL_604645\|2020-0 | ···················· | ···················· | ···················· | ···················· | ···················· | ···················· | |
| N | 2512 | hCoV-19/USA/CA-QDX-2345/2020\|EPI_ISL_604646\|2020-0 | ···················· | ···················· | ···················· | ···················· | ···················· | ···················· | |
| N | 2513 | hCoV-19/USA/CA-QDX-2346/2020\|EPI_ISL_604647\|2020-0 | ···················· | ···················· | ···················· | ···················· | ···················· | ···················· | |
| N | 2514 | hCoV-19/USA/CA-QDX-2349/2020\|EPI_ISL_604648\|2020-0 | ···················· | ···················· | ···················· | ···················· | ···················· | ···················· | |
| N | 2515 | hCoV-19/USA/CA-QDX-2350/2020\|EPI_ISL_604649\|2020-0 | ···················· | ···················· | ···················· | ···················· | ···················· | ···················· | |
| N | 2516 | hCoV-19/USA/CA-QDX-2351/2020\|EPI_ISL_604650\|2020-0 | ···················· | ···················· | ···················· | ···················· | ···················· | ···················· | |
| N | 2517 | hCoV-19/USA/CA-QDX-2353/2020\|EPI_ISL_604651\|2020-0 | ···················· | ···················· | ···················· | ···················· | ···················· | ···················· | |
| N | 2518 | hCoV-19/USA/CA-QDX-2355/2020\|EPI_ISL_604652\|2020-0 | ···················· | ···················· | ···················· | ···················· | ···················· | ···················· | |
| N | 2519 | hCoV-19/USA/CA-QDX-2357/2020\|EPI_ISL_604653\|2020-0 | ···················· | ···················· | ···················· | ···················· | ···················· | ···················· | |
| N | 2520 | hCoV-19/USA/CA-QDX-2358/2020\|EPI_ISL_604654\|2020-0 | ···················· | ···················· | ···················· | ···················· | ···················· | ···················· | |
| N | 2521 | hCoV-19/USA/CA-QDX-2361/2020\|EPI_ISL_604655\|2020-0 | ···················· | ···················· | ···················· | ···················· | ···················· | ···················· | |
| N | 2522 | hCoV-19/USA/CA-QDX-2362/2020\|EPI_ISL_604656\|2020-0 | ···················· | ···················· | ···················· | ···················· | ···················· | ···················· | |
| N | 2523 | hCoV-19/USA/CA-QDX-2371/2020\|EPI_ISL_604657\|2020-0 | ···················· | ···················· | ···················· | ···················· | ···················· | ···················· | |
| N | 2524 | hCoV-19/USA/CA-QDX-2374/2020\|EPI_ISL_604658\|2020-0 | ···················· | ···················· | ···················· | ···················· | ···················· | ···················· | |
| N | 2525 | hCoV-19/USA/CA-QDX-2375/2020\|EPI_ISL_604659\|2020-0 | ···················· | ···················· | ···················· | ···················· | ···················· | ···················· | |
| N | 2526 | hCoV-19/USA/CA-QDX-2381/2020\|EPI_ISL_604660\|2020-0 | ···················· | ···················· | ···················· | ···················· | ···················· | ···················· | |
| N | 2527 | hCoV-19/USA/CA-QDX-2382/2020\|EPI_ISL_604661\|2020-0 | ···················· | ···················· | ···················· | ···················· | ···················· | ···················· | |
| N | 2528 | hCoV-19/USA/CA-QDX-2383/2020\|EPI_ISL_604662\|2020-0 | ···················· | ···················· | ···················· | ···················· | ···················· | ···················· | |
| N | 2529 | hCoV-19/USA/CA-QDX-2385/2020\|EPI_ISL_604663\|2020-0 | ···················· | ···················· | ···················· | ···················· | ···················· | ···················· | |
| N | 2530 | hCoV-19/USA/CA-QDX-2387/2020\|EPI_ISL_604664\|2020-0 | ···················· | ···················· | ···················· | ···················· | ···················· | ···················· | |
| N | 2531 | hCoV-19/USA/CA-QDX-2390/2020\|EPI_ISL_604665\|2020-0 | ···················· | ···················· | ···················· | ···················· | ···················· | ···················· | |
| N | 2532 | hCoV-19/USA/CA-QDX-2392/2020\|EPI_ISL_604666\|2020-0 | ···················· | ···················· | ···················· | ···················· | ···················· | ···················· | |
| N | 2533 | hCoV-19/USA/CA-QDX-2393/2020\|EPI_ISL_604667\|2020-0 | ···················· | ···················· | ···················· | ···················· | ···················· | ···················· | |
| N | 2534 | hCoV-19/USA/CA-QDX-2394/2020\|EPI_ISL_604668\|2020-0 | ···················· | ···················· | ···················· | ···················· | ···················· | ···················· | |
| N | 2535 | hCoV-19/USA/CA-QDX-2395/2020\|EPI_ISL_604669\|2020-0 | ···················· | ···················· | ···················· | ···················· | ···················· | ···················· | |
| N | 2536 | hCoV-19/USA/CA-QDX-2397/2020\|EPI_ISL_604671\|2020-0 | ···················· | ···················· | ···················· | ···················· | ···················· | ···················· | |
| N | 2537 | hCoV-19/USA/CA-QDX-2398/2020\|EPI_ISL_604672\|2020-0 | ···················· | ···················· | ···················· | ···················· | ···················· | ···················· | |
| N | 2538 | hCoV-19/USA/CA-QDX-2405/2020\|EPI_ISL_604673\|2020-0 | ···················· | ···················· | ···················· | ···················· | ···················· | ···················· | |
| N | 2539 | hCoV-19/USA/CA-QDX-2406/2020\|EPI_ISL_604674\|2020-0 | ···················· | ···················· | ···················· | ···················· | ···················· | ···················· | |
| N | 2540 | hCoV-19/USA/CA-QDX-2408/2020\|EPI_ISL_604675\|2020-0 | ···················· | ···················· | ···················· | ···················· | ···················· | ···················· | |
| N | 2541 | hCoV-19/USA/CA-QDX-2410/2020\|EPI_ISL_604676\|2020-0 | ···················· | ···················· | ···················· | ···················· | ···················· | ···················· | |
| N | 2542 | hCoV-19/USA/CA-QDX-2411/2020\|EPI_ISL_604677\|2020-0 | ···················· | ···················· | ···················· | ···················· | ···················· | ···················· | |
| N | 2543 | hCoV-19/USA/CA-QDX-2412/2020\|EPI_ISL_604678\|2020-0 | ···················· | ···················· | ···················· | ···················· | ···················· | ···················· | |
| N | 2544 | hCoV-19/USA/CA-QDX-2413/2020\|EPI_ISL_604679\|2020-0 | ···················· | ···················· | ···················· | ···················· | ···················· | ···················· | |
| N | 2545 | hCoV-19/USA/MI-QDX-2360/2020\|EPI_ISL_604700\|2020-0 | ···················· | ···················· | ···················· | ···················· | ···················· | ···················· | |
| N | 2546 | hCoV-19/USA/MI-QDX-2372/2020\|EPI_ISL_604701\|2020-0 | ···················· | ···················· | ···················· | ···················· | ···················· | ···················· | |
| N | 2547 | hCoV-19/USA/MI-QDX-2399/2020\|EPI_ISL_604702\|2020-0 | ···················· | ···················· | ···················· | ···················· | ···················· | ···················· | |

N 2548 hCoV-19/USA/NJ-QDX-2400/2020|EPI_ISL_604703|2020-0 ···················· ···················· ····················· ···················· ···················· ····················
N 2549 hCoV-19/USA/NJ-QDX-2404/2020|EPI_ISL_604704|2020-0 ···················· ···················· ····················· ···················· ···················· ····················
N 2550 hCoV-19/USA/NY-QDX-2330/2020|EPI_ISL_604705|2020-0 ···················· ···················· ····················· ···················· ···················· ····················
N 2551 hCoV-19/USA/NY-QDX-2352/2020|EPI_ISL_604706|2020-0 ···················· ···················· ····················· ···················· ···················· ····················
N 2552 hCoV-19/USA/NY-QDX-2363/2020|EPI_ISL_604707|2020-0 ···················· ···················· ····················· ···················· ···················· ····················
N 2553 hCoV-19/USA/NY-QDX-2364/2020|EPI_ISL_604708|2020-0 ···················· ···················· ····················· ···················· ···················· ····················
N 2554 hCoV-19/USA/NY-QDX-2365/2020|EPI_ISL_604709|2020-0 ···················· ···················· ····················· ···················· ···················· ····················
N 2555 hCoV-19/USA/NY-QDX-2366/2020|EPI_ISL_604710|2020-0 ···················· ···················· ····················· ···················· ···················· ····················
N 2556 hCoV-19/USA/NY-QDX-2370/2020|EPI_ISL_604711|2020-0 ···················· ···················· ····················· ···················· ···················· ····················
N 2557 hCoV-19/USA/NY-QDX-2373/2020|EPI_ISL_604712|2020-0 ···················· ···················· ····················· ···················· ···················· ····················
N 2558 hCoV-19/USA/NY-QDX-2380/2020|EPI_ISL_604713|2020-0 ···················· ···················· ····················· ···················· ···················· ····················
N 2559 hCoV-19/USA/NY-QDX-2386/2020|EPI_ISL_604714|2020-0 ···················· ···················· ····················· ···················· ···················· ····················
N 2560 hCoV-19/USA/NY-QDX-2389/2020|EPI_ISL_604715|2020-0 ···················· ···················· ····················· ···················· ···················· ····················
N 2561 hCoV-19/England/ALDP-A5225A/2020|EPI_ISL_598248|20 ···················· ···················· ····················· ···················· ···················· ····················
N 2562 hCoV-19/England/MILK-A47944/2020|EPI_ISL_598251|20 ···················· ···················· ····················· ···················· ···················· ····················
N 2563 hCoV-19/England/MILK-A476A7/2020|EPI_ISL_598252|20 ···················· ·A·················· ····················· ···················· ···················· ····················
N 2564 hCoV-19/England/MILK-A47CC3/2020|EPI_ISL_598253|20 ···················· ···················· ····················· ···················· ···················· ····················
N 2565 hCoV-19/England/MILK-A4716A/2020|EPI_ISL_598254|20 ···················· ···················· ····················· ···················· ···················· ····················
N 2566 hCoV-19/England/MILK-A47F15/2020|EPI_ISL_598255|20 ···················· ···················· ····················· ···················· ···················· ····················
N 2567 hCoV-19/England/MILK-A46909/2020|EPI_ISL_598256|20 ···················· ···················· ····················· ···················· ···················· ····················
N 2568 hCoV-19/England/MILK-A478A1/2020|EPI_ISL_598257|20 ···················· ···················· ····················· ···················· ···················· ····················
N 2569 hCoV-19/England/MILK-A46B5E/2020|EPI_ISL_598258|20 ···················· ···················· ····················· ···················· ···················· ····················
N 2570 hCoV-19/England/MILK-A47C0F/2020|EPI_ISL_598259|20 ···················· ···················· ····················· ···················· ···················· ····················
N 2571 hCoV-19/England/MILK-A47D0C/2020|EPI_ISL_598260|20 ···················· ···················· ····················· ···················· ···················· ····················
N 2572 hCoV-19/Wales/MILK-A47D57/2020|EPI_ISL_598261|2020 ···················· ···················· ····················· ···················· ···················· ····················
N 2573 hCoV-19/England/MILK-A473EC/2020|EPI_ISL_598262|20 ···················· ···················· ····················· ···················· ···················· ····················
N 2574 hCoV-19/England/MILK-A472B2/2020|EPI_ISL_598263|20 ···················· ···················· ····················· ···················· ···················· ····················
N 2575 hCoV-19/England/MILK-A46D2B/2020|EPI_ISL_598264|20 ···················· ···················· ····················· ···················· ···················· ····················
N 2576 hCoV-19/England/MILK-A46DC1/2020|EPI_ISL_598265|20 ···················· ···················· ····················· ···················· ······T············· ····················
N 2577 hCoV-19/England/MILK-A4764D/2020|EPI_ISL_598266|20 ···················· ···················· ····················· ···················· ···················· ····················
N 2578 hCoV-19/England/MILK-A47953/2020|EPI_ISL_598267|20 ···················· ···················· ····················· ···················· ···················· ····················
N 2579 hCoV-19/England/MILK-A47AF6/2020|EPI_ISL_598268|20 ···················· ···················· ····················· ···················· ···················· ····················
N 2580 hCoV-19/England/MILK-A47285/2020|EPI_ISL_598270|20 ···················· ···················· ····················· ···················· ···················· ····················
N 2581 hCoV-19/England/MILK-A46E0A/2020|EPI_ISL_598271|20 ···················· ···················· ····················· ···················· ···················· ····················
N 2582 hCoV-19/England/MILK-A46D85/2020|EPI_ISL_598272|20 ···················· ···················· ····················· ···················· ···················· ····················
N 2583 hCoV-19/England/MILK-A480A8/2020|EPI_ISL_598273|20 ···················· ···················· ····················· ···················· ···················· ····················
N 2584 hCoV-19/England/MILK-A48020/2020|EPI_ISL_598274|20 ···················· ···················· ····················· ···················· ······T············· ····················
N 2585 hCoV-19/England/MILK-A47EFA/2020|EPI_ISL_598275|20 ···················· ···················· ····················· ···················· ···················· ····················
N 2586 hCoV-19/England/MILK-A47328/2020|EPI_ISL_598276|20 ···················· ···················· ····················· ···················· ···················· ····················
N 2587 hCoV-19/England/MILK-A47267/2020|EPI_ISL_598277|20 ···················· ·A·················· ····················· ···················· ···················· ····················
N 2588 hCoV-19/England/MILK-A4730A/2020|EPI_ISL_598278|20 ···················· ···················· ····················· ···················· ···················· ····················
N 2589 hCoV-19/Wales/MILK-A4770E/2020|EPI_ISL_598279|2020 ···················· ···················· ····················· ···················· ···················· ····················
N 2590 hCoV-19/England/MILK-A47B8A/2020|EPI_ISL_598280|20 ···················· ···················· ····················· ···················· ···················· ····················
N 2591 hCoV-19/England/MILK-A46F52/2020|EPI_ISL_598281|20 ···················· ···················· ····················· ···················· ···················· ····················
N 2592 hCoV-19/England/MILK-A479BD/2020|EPI_ISL_598282|20 ···················· ···················· ····················· ···················· ······T············· ····················
N 2593 hCoV-19/England/MILK-A47DDF/2020|EPI_ISL_598284|20 ···················· ···················· ····················· ···················· ···················· ····················
N 2594 hCoV-19/England/MILK-A473BF/2020|EPI_ISL_598286|20 ···················· ···················· ····················· ···················· ···················· ····················
N 2595 hCoV-19/England/MILK-A47C78/2020|EPI_ISL_598287|20 ···················· ···················· ····················· ···················· ···················· ····················
N 2596 hCoV-19/England/MILK-A47E36/2020|EPI_ISL_598288|20 ···················· ·A·················· ····················· ···················· ···················· ····················
N 2597 hCoV-19/England/MILK-A47B02/2020|EPI_ISL_598290|20 ···················· ·A·················· ····················· ···················· ···················· ···A················
N 2598 hCoV-19/England/MILK-A47FAC/2020|EPI_ISL_598291|20 ···················· ···················· ····················· ···················· ···················· ····················
N 2599 hCoV-19/England/MILK-A477C2/2020|EPI_ISL_598292|20 ···················· ···················· ····················· ···················· ···················· ····················
N 2600 hCoV-19/England/MILK-A47E27/2020|EPI_ISL_598293|20 ···················· ·A·················· ····················· ···················· ···················· ····················
N 2601 hCoV-19/England/MILK-A4759B/2020|EPI_ISL_598294|20 ···················· ···················· ····················· ···················· ···················· ····················
N 2602 hCoV-19/England/MILK-A474CB/2020|EPI_ISL_598295|20 ···················· ···················· ····················· ···················· ···················· ····················
N 2603 hCoV-19/England/MILK-A47F42/2020|EPI_ISL_598296|20 ···················· ···················· ····················· ···················· ···················· ····················
N 2604 hCoV-19/England/MILK-A46A24/2020|EPI_ISL_598297|20 ···················· ···················· ····················· ···················· ···················· ····················
N 2605 hCoV-19/England/MILK-A47F51/2020|EPI_ISL_598298|20 ···················· ···················· ····················· ···················· ···················· ····················
N 2606 hCoV-19/England/MILK-A46FE9/2020|EPI_ISL_598299|20 ···················· ···················· ····················· ···················· ···················· ····················
N 2607 hCoV-19/England/MILK-A47F24/2020|EPI_ISL_598303|20 ···················· ···················· ····················· ···················· ······T············· ····················
N 2608 hCoV-19/England/MILK-A46A7F/2020|EPI_ISL_598310|20 ···················· ···················· ····················· ···················· ···················· ····················
N 2609 hCoV-19/England/MILK-A471A6/2020|EPI_ISL_598311|20 ···················· ···················· ····················· ···················· ···················· ····················
N 2610 hCoV-19/England/MILK-A46B12/2020|EPI_ISL_598312|20 ···················· ···················· ····················· ···················· ···················· ····················
N 2611 hCoV-19/England/MILK-A46FDA/2020|EPI_ISL_598313|20 ···················· ···················· ····················· ···················· ······T············· ····················
N 2612 hCoV-19/England/QEUH-9DE363/2020|EPI_ISL_599579|20 ···················· ···················· ····················· ···················· ···················· ····················
N 2613 hCoV-19/England/QEUH-9DDB6C/2020|EPI_ISL_599580|20 ···················· ···················· ····················· ···················· ···················· ····················
N 2614 hCoV-19/England/QEUH-9DD70E/2020|EPI_ISL_599581|20 ···················· ·A·················· ····················· ···················· ···················· ····················
N 2615 hCoV-19/England/QEUH-9DE2B1/2020|EPI_ISL_599582|20 ···················· ···················· ····················· ···················· ···················· ····················
N 2616 hCoV-19/England/QEUH-9DDEAF/2020|EPI_ISL_599583|20 ···················· ···················· ····················· ···················· ···················· ····················
N 2617 hCoV-19/England/QEUH-9DE327/2020|EPI_ISL_599584|20 ···················· ·A·················· ····················· ···················· ···················· ···A················
N 2618 hCoV-19/England/QEUH-9DDB5D/2020|EPI_ISL_599585|20 ···················· ···················· ····················· ···················· ···················· ····················
N 2619 hCoV-19/England/QEUH-9DD980/2020|EPI_ISL_599586|20 ···················· ···················· ····················· ···················· ···················· ····················
N 2620 hCoV-19/Scotland/QEUH-9DD531/2020|EPI_ISL_599588|2 ···················· ···················· ····················· ···················· ···················· ····················
N 2621 hCoV-19/England/QEUH-9DD8FC/2020|EPI_ISL_599589|20 ···················· ···················· ····················· ···················· ···················· ····················
N 2622 hCoV-19/England/QEUH-9DE0F3/2020|EPI_ISL_599591|20 ···················· ···················· ····················· ···················· ···················· ····················
N 2623 hCoV-19/Scotland/QEUH-9DD3CE/2020|EPI_ISL_599593|2 ···················· ···················· ····················· ···················· ···················· ····················
N 2624 hCoV-19/England/QEUH-9DDCC3/2020|EPI_ISL_599594|20 ···················· ······C············· ····················· ···················· ···················· ····················
N 2625 hCoV-19/Scotland/QEUH-9DD391/2020|EPI_ISL_599596|2 ···················· ···················· ····················· ···················· ···················· ····················
N 2626 hCoV-19/Scotland/QEUH-9DD276/2020|EPI_ISL_599597|2 ···················· ···················· ····················· ···················· ···················· ····················
N 2627 hCoV-19/England/QEUH-9DD962/2020|EPI_ISL_599598|20 ···················· ···················· ····················· ···················· ···················· ····················

| | | | | | | | |
|---|---|---|---|---|---|---|---|
| N | 2628 hCoV-19/England/QEUH-9DCFCB/2020\|EPI_ISL_599642\|20 | ···················· | ···················· | ···················· | ···················· | ···················· | ···················· |
| N | 2629 hCoV-19/England/QEUH-9DE3CD/2020\|EPI_ISL_599643\|20 | ···················· | ···················· | ···················· | ···················· | ···················· | ···················· |
| N | 2630 hCoV-19/England/QEUH-9DE2EE/2020\|EPI_ISL_599644\|20 | ···················· | ···················· | ···················· | ···················· | ···················· | ···················· |
| N | 2631 hCoV-19/Scotland/QEUH-9CE69A/2020\|EPI_ISL_602010\|2 | ···················· | ···················· | ···················· | ···················· | ···················· | ···················· |
| N | 2632 hCoV-19/Scotland/QEUH-9CE2F0/2020\|EPI_ISL_602011\|2 | ···················· | ···················· | ···················· | ···················· | ···················· | ···················· |
| N | 2633 hCoV-19/Scotland/QEUH-9CF72D/2020\|EPI_ISL_602020\|2 | ···················· | ···················· | ···················· | ···················· | ·····T·············· | ···················· |
| N | 2634 hCoV-19/Scotland/QEUH-9CE2C3/2020\|EPI_ISL_602021\|2 | ···················· | ···················· | ···················· | ···················· | ···················· | ···················· |
| N | 2635 hCoV-19/Scotland/QEUH-9CE515/2020\|EPI_ISL_602022\|2 | ···················· | ···················· | ···················· | ···················· | ···················· | ···················· |
| N | 2636 hCoV-19/Scotland/QEUH-9CEE0B/2020\|EPI_ISL_602023\|2 | ···················· | ···················· | ···················· | ···················· | ···················· | ···················· |
| N | 2637 hCoV-19/Scotland/QEUH-9CF523/2020\|EPI_ISL_602024\|2 | ···················· | ···················· | ···················· | ···················· | ···················· | ···················· |
| N | 2638 hCoV-19/Scotland/QEUH-9CE418/2020\|EPI_ISL_602025\|2 | ···················· | ···················· | ···················· | ···················· | ···················· | ···················· |
| N | 2639 hCoV-19/England/MILK-9C2441/2020\|EPI_ISL_602027\|20 | ···················· | ···················· | ···················· | ···················· | ···················· | ···················· |
| N | 2640 hCoV-19/England/MILK-9C4333/2020\|EPI_ISL_602029\|20 | ···················· | ···················· | ···················· | ···················· | ···················· | ···················· |
| N | 2641 hCoV-19/England/QEUH-9C7F1E/2020\|EPI_ISL_602030\|20 | ···················· | ···················· | ···················· | ···················· | ···················· | ···················· |
| N | 2642 hCoV-19/England/QEUH-9C7F0F/2020\|EPI_ISL_602031\|20 | ···················· | ···················· | ···················· | ···················· | ···················· | ···················· |
| N | 2643 hCoV-19/England/QEUH-9C932F/2020\|EPI_ISL_602033\|20 | ···················· | ···················· | ···················· | ···················· | ···················· | ···················· |
| N | 2644 hCoV-19/England/QEUH-9C985D/2020\|EPI_ISL_602034\|20 | ···················· | ···················· | ···················· | ···················· | ···················· | ···················· |
| N | 2645 hCoV-19/England/QEUH-9CA69E/2020\|EPI_ISL_602035\|20 | ···················· | ···················· | ···················· | ···················· | ···················· | ···················· |
| N | 2646 hCoV-19/England/QEUH-9C9A57/2020\|EPI_ISL_602036\|20 | ···················· | ···················· | ···················· | ···················· | ···················· | ···················· |
| N | 2647 hCoV-19/England/QEUH-9C9899/2020\|EPI_ISL_602037\|20 | ···················· | ···················· | ···················· | ···················· | ···················· | ···················· |
| N | 2648 hCoV-19/England/QEUH-9CA889/2020\|EPI_ISL_602038\|20 | ···················· | ···················· | ···················· | ···················· | ···················· | ···················· |
| N | 2649 hCoV-19/England/QEUH-9CA6BC/2020\|EPI_ISL_602039\|20 | ···················· | ···················· | ···················· | ···················· | ···················· | ···················· |
| N | 2650 hCoV-19/England/QEUH-9CA6AD/2020\|EPI_ISL_602040\|20 | ···················· | ···················· | ···················· | ···················· | ···················· | ···················· |
| N | 2651 hCoV-19/England/QEUH-9C93B6/2020\|EPI_ISL_602041\|20 | ···················· | ···················· | ···················· | ···················· | ···················· | ···················· |
| N | 2652 hCoV-19/England/QEUH-9CA0DC/2020\|EPI_ISL_602042\|20 | ···················· | ···················· | ···················· | ···················· | ···················· | ···················· |
| N | 2653 hCoV-19/Scotland/QEUH-9B510E/2020\|EPI_ISL_602043\|2 | ···················· | ···················· | ···················· | ···················· | ···················· | ···················· |
| N | 2654 hCoV-19/Scotland/QEUH-9B5177/2020\|EPI_ISL_602044\|2 | ···················· | ···················· | ···················· | ···················· | ···················· | ···················· |
| N | 2655 hCoV-19/Scotland/QEUH-9B4A7D/2020\|EPI_ISL_602045\|2 | ···················· | ···················· | ···················· | ···················· | ···················· | ···················· |
| N | 2656 hCoV-19/Scotland/QEUH-9B54BA/2020\|EPI_ISL_602046\|2 | ···················· | ···················· | ···················· | ···················· | ···················· | ···················· |
| N | 2657 hCoV-19/Scotland/QEUH-9B4EF9/2020\|EPI_ISL_602047\|2 | ···················· | ···················· | ···················· | ···················· | ···················· | ···················· |
| N | 2658 hCoV-19/Scotland/QEUH-9B4ECC/2020\|EPI_ISL_602048\|2 | ···················· | ···················· | ···················· | ···················· | ···················· | ···················· |
| N | 2659 hCoV-19/Scotland/QEUH-9B54D8/2020\|EPI_ISL_602049\|2 | ···················· | ···················· | ···················· | ···················· | ···················· | ···················· |
| N | 2660 hCoV-19/Scotland/QEUH-9B4925/2020\|EPI_ISL_602050\|2 | ···················· | ···················· | ···················· | ···················· | ···················· | ···················· |
| N | 2661 hCoV-19/Scotland/QEUH-9B4AF5/2020\|EPI_ISL_602051\|2 | ···················· | ···················· | ···················· | ···················· | ···················· | ···················· |
| N | 2662 hCoV-19/Scotland/QEUH-9B5089/2020\|EPI_ISL_602052\|2 | ···················· | ···················· | ···················· | ···················· | ···················· | ···················· |
| N | 2663 hCoV-19/Scotland/QEUH-9B546F/2020\|EPI_ISL_602053\|2 | ···················· | ···················· | ···················· | ···················· | ···················· | ···················· |
| N | 2664 hCoV-19/Scotland/QEUH-9B4BD4/2020\|EPI_ISL_602054\|2 | ···················· | ···················· | ···················· | ···················· | ···················· | ···················· |
| N | 2665 hCoV-19/Scotland/QEUH-9B4BB6/2020\|EPI_ISL_602055\|2 | ···················· | ···················· | ···················· | ···················· | ···················· | ···················· |
| N | 2666 hCoV-19/Scotland/QEUH-9B4AD7/2020\|EPI_ISL_602056\|2 | ···················· | ···················· | ···················· | ···················· | ···················· | ···················· |
| N | 2667 hCoV-19/Scotland/QEUH-9B50D4/2020\|EPI_ISL_602057\|2 | ···················· | ···················· | ···················· | ···················· | ···················· | ···················· |
| N | 2668 hCoV-19/Scotland/QEUH-9B4AC8/2020\|EPI_ISL_602058\|2 | ···················· | ···················· | ···················· | ···················· | ···················· | ···················· |
| N | 2669 hCoV-19/Scotland/QEUH-9B461F/2020\|EPI_ISL_602059\|2 | ···················· | ···················· | ···················· | ···················· | ···················· | ···················· |
| N | 2670 hCoV-19/Scotland/QEUH-9B45D6/2020\|EPI_ISL_602060\|2 | ···················· | ···················· | ···················· | ···················· | ···················· | ···················· |
| N | 2671 hCoV-19/Scotland/QEUH-9B45B8/2020\|EPI_ISL_602061\|2 | ···················· | ···················· | ···················· | ···················· | ···················· | ···················· |
| N | 2672 hCoV-19/Scotland/QEUH-9B50F2/2020\|EPI_ISL_602062\|2 | ···················· | ···················· | ···················· | ···················· | ···················· | ···················· |
| N | 2673 hCoV-19/Scotland/QEUH-9B4FC9/2020\|EPI_ISL_602063\|2 | ···················· | ···················· | ···················· | ···················· | ···················· | ···················· |
| N | 2674 hCoV-19/Scotland/QEUH-9B54F6/2020\|EPI_ISL_602064\|2 | ···················· | ···················· | ···················· | ···················· | ···················· | ···················· |
| N | 2675 hCoV-19/Scotland/QEUH-9B4CD1/2020\|EPI_ISL_602065\|2 | ···················· | ···················· | ···················· | ···················· | ···················· | ···················· |
| N | 2676 hCoV-19/Scotland/QEUH-9B4B01/2020\|EPI_ISL_602066\|2 | ···················· | ·A·················· | ···················· | ···················· | ···················· | ···················· |
| N | 2677 hCoV-19/Scotland/QEUH-9B513B/2020\|EPI_ISL_602067\|2 | ···················· | ···················· | ···················· | ···················· | ···················· | ···················· |
| N | 2678 hCoV-19/Scotland/QEUH-9B4AE6/2020\|EPI_ISL_602068\|2 | ···················· | ···················· | ···················· | ···················· | ···················· | ···················· |
| N | 2679 hCoV-19/Scotland/QEUH-9B548D/2020\|EPI_ISL_602069\|2 | ···················· | ···················· | ···················· | ···················· | ···················· | ···················· |
| N | 2680 hCoV-19/Scotland/QEUH-9B4EDB/2020\|EPI_ISL_602070\|2 | ···················· | ···················· | ···················· | ···················· | ···················· | ···················· |
| N | 2681 hCoV-19/Scotland/QEUH-9B5450/2020\|EPI_ISL_602071\|2 | ···················· | ···················· | ···················· | ···················· | ···················· | ···················· |
| N | 2682 hCoV-19/Scotland/QEUH-9B52CF/2020\|EPI_ISL_602072\|2 | ···················· | ···················· | ···················· | ···················· | ···················· | ···················· |
| N | 2683 hCoV-19/Scotland/QEUH-9B4E44/2020\|EPI_ISL_602073\|2 | ···················· | ···················· | ···················· | ···················· | ···················· | ···················· |
| N | 2684 hCoV-19/Scotland/QEUH-9B45F4/2020\|EPI_ISL_602074\|2 | ···················· | ···················· | ···················· | ···················· | ···················· | ···················· |
| N | 2685 hCoV-19/Scotland/QEUH-9B52ED/2020\|EPI_ISL_602075\|2 | ···················· | ···················· | ···················· | ···················· | ···················· | ···················· |
| N | 2686 hCoV-19/Scotland/QEUH-9B4F32/2020\|EPI_ISL_602076\|2 | ···················· | ···················· | ···················· | ···················· | ···················· | ···················· |
| N | 2687 hCoV-19/Scotland/QEUH-9B4F23/2020\|EPI_ISL_602078\|2 | ···················· | ···················· | ···················· | ···················· | ···················· | ···················· |
| N | 2688 hCoV-19/Scotland/QEUH-9B4F05/2020\|EPI_ISL_602079\|2 | ···················· | ···················· | ···················· | ···················· | ···················· | ···················· |
| N | 2689 hCoV-19/Scotland/QEUH-9B547E/2020\|EPI_ISL_602080\|2 | ···················· | ···················· | ···················· | ···················· | ···················· | ···················· |
| N | 2690 hCoV-19/Scotland/QEUH-9B4CE0/2020\|EPI_ISL_602084\|2 | ···················· | ···················· | ···················· | ···················· | ···················· | ···················· |
| N | 2691 hCoV-19/Scotland/QEUH-9B5441/2020\|EPI_ISL_602085\|2 | ···················· | ···················· | ···················· | ···················· | ···················· | ···················· |
| N | 2692 hCoV-19/Scotland/QEUH-9B4CA4/2020\|EPI_ISL_602086\|2 | ···················· | ···················· | ···················· | ···················· | ···················· | ···················· |
| N | 2693 hCoV-19/Scotland/QEUH-9B4F8D/2020\|EPI_ISL_602087\|2 | ···················· | ···················· | ···················· | ···················· | ···················· | ···················· |
| N | 2694 hCoV-19/Scotland/QEUH-9B4F6F/2020\|EPI_ISL_602088\|2 | ···················· | ···················· | ···················· | ···················· | ···················· | ···················· |
| N | 2695 hCoV-19/Scotland/QEUH-9B73F7/2020\|EPI_ISL_602089\|2 | ···················· | ···················· | ···················· | ···················· | ···················· | ···················· |
| N | 2696 hCoV-19/Scotland/QEUH-9B73E8/2020\|EPI_ISL_602093\|2 | ···················· | ···················· | ···················· | ···················· | ···················· | ···················· |
| N | 2697 hCoV-19/Scotland/QEUH-9B6E7F/2020\|EPI_ISL_602094\|2 | ···················· | ···················· | ···················· | ···················· | ···················· | ···················· |
| N | 2698 hCoV-19/Scotland/QEUH-9B7E7E/2020\|EPI_ISL_602095\|2 | ···················· | ···················· | ···················· | ···················· | ···················· | ···················· |
| N | 2699 hCoV-19/Scotland/QEUH-9B7AE3/2020\|EPI_ISL_602096\|2 | ···················· | ···················· | ···················· | ···················· | ···················· | ···················· |
| N | 2700 hCoV-19/Scotland/QEUH-9B7755/2020\|EPI_ISL_602097\|2 | ···················· | ···················· | ···················· | ···················· | ···················· | ···················· |
| N | 2701 hCoV-19/Scotland/QEUH-9B7764/2020\|EPI_ISL_602098\|2 | ···················· | ···················· | ···················· | ···················· | ···················· | ···················· |
| N | 2702 hCoV-19/England/QEUH-9B7078/2020\|EPI_ISL_602099\|20 | ···················· | ···················· | ···················· | ···················· | ···················· | ···················· |
| N | 2703 hCoV-19/Switzerland/VD-ETHZ-300268/2020\|EPI_ISL_60 | ···················· | ···················· | ···················· | ···················· | ···················· | ···················· |
| N | 2704 hCoV-19/Switzerland/VD-ETHZ-300269/2020\|EPI_ISL_60 | ···················· | ···················· | ···················· | ···················· | ···················· | ···················· |
| N | 2705 hCoV-19/Switzerland/VD-ETHZ-300270/2020\|EPI_ISL_60 | ···················· | ···················· | ···················· | ···················· | ···················· | ···················· |
| N | 2706 hCoV-19/Switzerland/VD-ETHZ-300271/2020\|EPI_ISL_60 | ···················· | ···················· | ···················· | ···················· | ···················· | ···················· |
| N | 2707 hCoV-19/Switzerland/VD-ETHZ-300273/2020\|EPI_ISL_60 | ···················· | ···················· | ···················· | ···················· | ···················· | ···················· |

| | | | | | | | |
|---|---|---|---|---|---|---|---|
| N | 2708 hCoV-19/Switzerland/VD-ETHZ-300274/2020\|EPI_ISL_60 | ···················· | ···················· | ···················· | ···················· | ···················· | ···················· |
| N | 2709 hCoV-19/Switzerland/VD-ETHZ-300318/2020\|EPI_ISL_60 | ···················· | ···················· | ···················· | ···················· | ···················· | ···················· |
| N | 2710 hCoV-19/Switzerland/VD-ETHZ-300319/2020\|EPI_ISL_60 | ···················· | ···················· | ···················· | ···················· | ···················· | ···················· |
| N | 2711 hCoV-19/Switzerland/ZH-ETHZ-300320/2020\|EPI_ISL_60 | ···················· | ···················· | ···················· | ···················· | ···················· | ···················· |
| N | 2712 hCoV-19/Switzerland/SG-ETHZ-300321/2020\|EPI_ISL_60 | ···················· | ···················· | ···················· | ···················· | ···················· | ···················· |
| N | 2713 hCoV-19/Switzerland/ZH-ETHZ-300322/2020\|EPI_ISL_60 | ···················· | ···················· | ···················· | ···················· | ···················· | ···················· |
| N | 2714 hCoV-19/Switzerland/BL-ETHZ-300323/2020\|EPI_ISL_60 | ···················· | ·A·················· | ···················· | ···················· | ···················· | ···················· |
| N | 2715 hCoV-19/Switzerland/BE-ETHZ-300325/2020\|EPI_ISL_60 | ···················· | ···················· | ···················· | ···················· | ···················· | ···················· |
| N | 2716 hCoV-19/Switzerland/ZH-ETHZ-300389/2020\|EPI_ISL_60 | ···················· | ···················· | ···················· | ···················· | ···················· | ···················· |
| N | 2717 hCoV-19/Switzerland/BE-ETHZ-300392/2020\|EPI_ISL_60 | ···················· | ···················· | ···················· | ···················· | ···················· | ···················· |
| N | 2718 hCoV-19/Switzerland/BE-ETHZ-300393/2020\|EPI_ISL_60 | ···················· | ···················· | ···················· | ···················· | ···················· | ···················· |
| N | 2719 hCoV-19/Switzerland/BE-ETHZ-300394/2020\|EPI_ISL_60 | ···················· | ···················· | ···················· | ···················· | ···················· | ···················· |
| N | 2720 hCoV-19/Switzerland/BE-ETHZ-300395/2020\|EPI_ISL_60 | ···················· | ···················· | ···················· | ···················· | ···················· | ···················· |
| N | 2721 hCoV-19/Switzerland/BE-ETHZ-300396/2020\|EPI_ISL_60 | ···················· | ···················· | ···················· | ···················· | ···················· | ···················· |
| N | 2722 hCoV-19/Switzerland/BE-ETHZ-300397/2020\|EPI_ISL_60 | ···················· | ···················· | ···················· | ···················· | ···················· | ···················· |
| N | 2723 hCoV-19/Switzerland/ZH-ETHZ-300398/2020\|EPI_ISL_60 | ···················· | ···················· | ···················· | ···················· | ···················· | ···················· |
| N | 2724 hCoV-19/Switzerland/ZH-ETHZ-300399/2020\|EPI_ISL_60 | ···················· | ···················· | ···················· | ···················· | ···················· | ···················· |
| N | 2725 hCoV-19/Switzerland/ZH-ETHZ-300401/2020\|EPI_ISL_60 | ···················· | ···················· | ···················· | ···················· | ···················· | ···················· |
| N | 2726 hCoV-19/Switzerland/BL-ETHZ-300403/2020\|EPI_ISL_60 | ···················· | ···················· | ···················· | ···················· | ···················· | ···················· |
| N | 2727 hCoV-19/Switzerland/ZH-ETHZ-300404/2020\|EPI_ISL_60 | ···················· | ···················· | ···················· | ···················· | ···················· | ···················· |
| N | 2728 hCoV-19/Switzerland/BE-ETHZ-300405/2020\|EPI_ISL_60 | ···················· | ···················· | ···················· | ···················· | ···················· | ···················· |
| N | 2729 hCoV-19/Switzerland/ZH-ETHZ-300406/2020\|EPI_ISL_60 | ···················· | ···················· | ···················· | ···················· | ···················· | ···················· |
| N | 2730 hCoV-19/Switzerland/GE-ETHZ-300408/2020\|EPI_ISL_60 | ···T················ | ·A·················· | ···················· | ···················· | ···················· | ···················· |
| N | 2731 hCoV-19/Switzerland/VS-ETHZ-300411/2020\|EPI_ISL_60 | ···················· | ···················· | ···················· | ···················· | ···················· | ···················· |
| N | 2732 hCoV-19/Switzerland/SO-ETHZ-300412/2020\|EPI_ISL_60 | ···················· | ···················· | ···················· | ···················· | ···········R········ | ···················· |
| N | 2733 hCoV-19/Switzerland/AG-ETHZ-300414/2020\|EPI_ISL_60 | ···················· | ···················· | ···················· | ···················· | ···················· | ···················· |
| N | 2734 hCoV-19/Switzerland/BE-ETHZ-300415/2020\|EPI_ISL_60 | ···················· | ···················· | ···················· | ···················· | ···················· | ···················· |
| N | 2735 hCoV-19/Switzerland/BE-ETHZ-300416/2020\|EPI_ISL_60 | ···················· | ···················· | ···················· | ···················· | ···················· | ···················· |
| N | 2736 hCoV-19/Switzerland/VD-ETHZ-300419/2020\|EPI_ISL_60 | ···················· | ···················· | ···················· | ···················· | ···················· | ···················· |
| N | 2737 hCoV-19/Switzerland/VD-ETHZ-300420/2020\|EPI_ISL_60 | ···················· | ···················· | ···················· | ···················· | ···················· | ···················· |
| N | 2738 hCoV-19/Switzerland/VD-ETHZ-300421/2020\|EPI_ISL_60 | ···················· | ···················· | ···················· | ···················· | ···················· | ···················· |
| N | 2739 hCoV-19/Switzerland/VD-ETHZ-300423/2020\|EPI_ISL_60 | ···················· | ···················· | ···················· | ···················· | ···················· | ···················· |
| N | 2740 hCoV-19/Switzerland/VD-ETHZ-300425/2020\|EPI_ISL_60 | ···················· | ···················· | ···················· | ···················· | ···················· | ···················· |
| N | 2741 hCoV-19/Switzerland/VD-ETHZ-300428/2020\|EPI_ISL_60 | ···················· | ···················· | ···················· | ···················· | ···················· | ···················· |
| N | 2742 hCoV-19/Switzerland/BE-ETHZ-300429/2020\|EPI_ISL_60 | ···················· | ···················· | ···················· | ···················· | ···················· | ···················· |
| N | 2743 hCoV-19/Switzerland/BE-ETHZ-300430/2020\|EPI_ISL_60 | ···················· | ···················· | ···················· | ···················· | ···················· | ···················· |
| N | 2744 hCoV-19/Switzerland/BE-ETHZ-300431/2020\|EPI_ISL_60 | ···················· | ···················· | ···················· | ···················· | ···················· | ···················· |
| N | 2745 hCoV-19/Switzerland/BE-ETHZ-300432/2020\|EPI_ISL_60 | ···················· | ···················· | ···················· | ···················· | ···················· | ···················· |
| N | 2746 hCoV-19/Switzerland/VD-ETHZ-300433/2020\|EPI_ISL_60 | ···················· | ···················· | ···················· | ···················· | ···················· | ···················· |
| N | 2747 hCoV-19/Switzerland/VD-ETHZ-300434/2020\|EPI_ISL_60 | ···················· | ···················· | ···················· | ···················· | ···················· | ···················· |
| N | 2748 hCoV-19/Switzerland/VS-ETHZ-300436/2020\|EPI_ISL_60 | ···················· | ···················· | ···················· | ···················· | ···················· | ···················· |
| N | 2749 hCoV-19/Switzerland/UR-ETHZ-300437/2020\|EPI_ISL_60 | ···················· | ···················· | ···················· | ···················· | ···················· | ···················· |
| N | 2750 hCoV-19/Switzerland/GE-ETHZ-300439/2020\|EPI_ISL_60 | ···················· | ···················· | ···················· | ···················· | ···················· | ···················· |
| N | 2751 hCoV-19/Switzerland/VS-ETHZ-300440/2020\|EPI_ISL_60 | ···················· | ···················· | ···················· | ···················· | ···················· | ···················· |
| N | 2752 hCoV-19/Switzerland/BE-ETHZ-300441/2020\|EPI_ISL_60 | ···················· | ···················· | ···················· | ···················· | ···················· | ···················· |
| N | 2753 hCoV-19/Switzerland/BE-ETHZ-300442/2020\|EPI_ISL_60 | ···················· | ···················· | ···················· | ···················· | ···················· | ···················· |
| N | 2754 hCoV-19/Switzerland/BE-ETHZ-300443/2020\|EPI_ISL_60 | ···················· | ···················· | ···················· | ···················· | ···················· | ···················· |
| N | 2755 hCoV-19/Switzerland/BE-ETHZ-300444/2020\|EPI_ISL_60 | ···················· | ···················· | ···················· | ···················· | ···················· | ···················· |
| N | 2756 hCoV-19/Switzerland/AG-ETHZ-300445/2020\|EPI_ISL_60 | ···················· | ···················· | ···················· | ···················· | ···················· | ···················· |
| N | 2757 hCoV-19/Switzerland/BE-ETHZ-300446/2020\|EPI_ISL_60 | ···················· | ···················· | ···················· | ···················· | ···················· | ···················· |
| N | 2758 hCoV-19/USA/UT-QDX-2253/2020\|EPI_ISL_604600\|2020-0 | ···················· | ···················· | ···················· | ···················· | ···················· | ···················· |
| N | 2759 hCoV-19/USA/UT-QDX-2257/2020\|EPI_ISL_604601\|2020-0 | ···················· | ···················· | ···················· | ···················· | ···················· | ···················· |
| N | 2760 hCoV-19/USA/UT-QDX-2262/2020\|EPI_ISL_604602\|2020-0 | ···················· | ···················· | ···················· | ···················· | ···················· | ···················· |
| N | 2761 hCoV-19/USA/UT-QDX-2264/2020\|EPI_ISL_604603\|2020-0 | ···················· | ···················· | ···················· | ···················· | ···················· | ···················· |
| N | 2762 hCoV-19/USA/UT-QDX-2270/2020\|EPI_ISL_604604\|2020-0 | ···················· | ···················· | ···················· | ···················· | ···················· | ···················· |
| N | 2763 hCoV-19/USA/UT-QDX-2281/2020\|EPI_ISL_604605\|2020-0 | ···················· | ···················· | ···················· | ···················· | ···················· | ···················· |
| N | 2764 hCoV-19/USA/UT-QDX-2285/2020\|EPI_ISL_604606\|2020-0 | ···················· | ···················· | ···················· | ···················· | ···················· | ···················· |
| N | 2765 hCoV-19/USA/UT-QDX-2286/2020\|EPI_ISL_604607\|2020-0 | ···················· | ···················· | ···················· | ···················· | ···················· | ···················· |
| N | 2766 hCoV-19/USA/UT-QDX-2287/2020\|EPI_ISL_604608\|2020-0 | ···················· | ···················· | ···················· | ···················· | ···················· | ···················· |
| N | 2767 hCoV-19/USA/UT-QDX-2289/2020\|EPI_ISL_604609\|2020-0 | ···················· | ···················· | ···················· | ···················· | ···················· | ···················· |
| N | 2768 hCoV-19/USA/UT-QDX-2290/2020\|EPI_ISL_604610\|2020-0 | ···················· | ···················· | ···················· | ···················· | ···················· | ···················· |
| N | 2769 hCoV-19/USA/UT-QDX-2295/2020\|EPI_ISL_604611\|2020-0 | ···················· | ···················· | ···················· | ···················· | ···················· | ···················· |
| N | 2770 hCoV-19/USA/UT-QDX-2296/2020\|EPI_ISL_604612\|2020-0 | ···················· | ···················· | ···················· | ···················· | ···················· | ···················· |
| N | 2771 hCoV-19/USA/UT-QDX-2297/2020\|EPI_ISL_604613\|2020-0 | ···················· | ···················· | ···················· | ···················· | ···················· | ···················· |
| N | 2772 hCoV-19/USA/UT-QDX-2300/2020\|EPI_ISL_604614\|2020-0 | ···················· | ···················· | ···················· | ···················· | ···················· | ···················· |
| N | 2773 hCoV-19/USA/UT-QDX-2301/2020\|EPI_ISL_604615\|2020-0 | ···················· | ···················· | ···················· | ···················· | ···················· | ···················· |
| N | 2774 hCoV-19/USA/UT-QDX-2302/2020\|EPI_ISL_604616\|2020-0 | ···················· | ···················· | ···················· | ···················· | ···················· | ···················· |
| N | 2775 hCoV-19/USA/UT-QDX-2305/2020\|EPI_ISL_604617\|2020-0 | ···················· | ···················· | ···················· | ···················· | ···················· | ···················· |
| N | 2776 hCoV-19/USA/UT-QDX-2307/2020\|EPI_ISL_604618\|2020-0 | ···················· | ···················· | ···················· | ···················· | ···················· | ···················· |
| N | 2777 hCoV-19/USA/UT-QDX-2308/2020\|EPI_ISL_604619\|2020-0 | ···················· | ···················· | ···················· | ···················· | ···················· | ···················· |
| N | 2778 hCoV-19/USA/UT-QDX-2313/2020\|EPI_ISL_604620\|2020-0 | ···················· | ···················· | ···················· | ···················· | ···················· | ···················· |
| N | 2779 hCoV-19/USA/VA-QDX-2211/2020\|EPI_ISL_604621\|2020-0 | ···················· | ···················· | ···················· | ···················· | ···················· | ···················· |
| N | 2780 hCoV-19/USA/WA-QDX-2241/2020\|EPI_ISL_604622\|2020-0 | ···················· | ···················· | ···················· | ···················· | ···················· | ···················· |
| N | 2781 hCoV-19/USA/WA-QDX-2242/2020\|EPI_ISL_604623\|2020-0 | ···················· | ···················· | ···················· | ···················· | ···················· | ···················· |
| N | 2782 hCoV-19/USA/WA-QDX-2276/2020\|EPI_ISL_604624\|2020-0 | ···················· | ···················· | ···················· | ···················· | ···················· | ···················· |
| N | 2783 hCoV-19/USA/AZ-QDX-2336/2020\|EPI_ISL_604626\|2020-0 | ···················· | ···················· | ···················· | ···················· | ···················· | ···················· |
| N | 2784 hCoV-19/USA/AZ-QDX-2384/2020\|EPI_ISL_604627\|2020-0 | ···················· | ···················· | ···················· | ···················· | ···················· | ···················· |
| N | 2785 hCoV-19/USA/CA-QDX-2317/2020\|EPI_ISL_604628\|2020-0 | ···················· | ···················· | ···················· | ···················· | ···················· | ···················· |
| N | 2786 hCoV-19/USA/CA-QDX-2318/2020\|EPI_ISL_604629\|2020-0 | ···················· | ···················· | ···················· | ···················· | ···················· | ···················· |
| N | 2787 hCoV-19/USA/CA-QDX-2320/2020\|EPI_ISL_604630\|2020-0 | ···················· | ···················· | ···················· | ···················· | ···················· | ···················· |

| | | | | | | | | |
|---|---|---|---|---|---|---|---|---|
| N | 2788 | hCoV-19/USA/CA-QDX-2323/2020\|EPI_ISL_604631\|2020-0 | ···················· | ···················· | ···················· | ···················· | ···················· | ···················· |
| N | 2789 | hCoV-19/USA/CA-QDX-2324/2020\|EPI_ISL_604632\|2020-0 | ···················· | ···················· | ···················· | ···················· | ···················· | ···················· |
| N | 2790 | hCoV-19/USA/CA-QDX-2325/2020\|EPI_ISL_604633\|2020-0 | ···················· | ···················· | ···················· | ···················· | ···················· | ···················· |
| N | 2791 | hCoV-19/USA/CA-QDX-2329/2020\|EPI_ISL_604634\|2020-0 | ···················· | ···················· | ···················· | ···················· | ···················· | ···················· |
| N | 2792 | hCoV-19/USA/CA-QDX-2331/2020\|EPI_ISL_604635\|2020-0 | ···················· | ···················· | ···················· | ···················· | ···················· | ···················· |
| N | 2793 | hCoV-19/USA/CA-QDX-2332/2020\|EPI_ISL_604636\|2020-0 | ···················· | ···················· | ···················· | ···················· | ···················· | ···················· |
| N | 2794 | hCoV-19/USA/CA-QDX-2334/2020\|EPI_ISL_604637\|2020-0 | ···················· | ···················· | ···················· | ···················· | ···················· | ···················· |
| N | 2795 | hCoV-19/USA/CA-QDX-2335/2020\|EPI_ISL_604638\|2020-0 | ···················· | ···················· | ···················· | ···················· | ···················· | ···················· |
| N | 2796 | hCoV-19/USA/CA-QDX-2396/2020\|EPI_ISL_604670\|2020-0 | ···················· | ···················· | ···················· | ···················· | ···················· | ···················· |
| N | 2797 | hCoV-19/USA/CA-QDX-2416/2020\|EPI_ISL_604680\|2020-0 | ···················· | ···················· | ···················· | ···················· | ···················· | ···················· |
| N | 2798 | hCoV-19/USA/CA-QDX-2418/2020\|EPI_ISL_604681\|2020-0 | ···················· | ···················· | ···················· | ···················· | ···················· | ···················· |
| N | 2799 | hCoV-19/USA/CA-QDX-2419/2020\|EPI_ISL_604682\|2020-0 | ···················· | ···················· | ···················· | ···················· | ···················· | ···················· |
| N | 2800 | hCoV-19/USA/CA-QDX-2424/2020\|EPI_ISL_604683\|2020-0 | ···················· | ···················· | ···················· | ···················· | ···················· | ···················· |
| N | 2801 | hCoV-19/USA/CO-QDX-2314/2020\|EPI_ISL_604684\|2020-0 | ···················· | ···················· | ···················· | ···················· | ···················· | ···················· |
| N | 2802 | hCoV-19/USA/CO-QDX-2322/2020\|EPI_ISL_604685\|2020-0 | ···················· | ···················· | ···················· | ···················· | ···················· | ···················· |
| N | 2803 | hCoV-19/USA/CO-QDX-2328/2020\|EPI_ISL_604686\|2020-0 | ···················· | ···················· | ···················· | ···················· | ···················· | ···················· |
| N | 2804 | hCoV-19/USA/CO-QDX-2348/2020\|EPI_ISL_604687\|2020-0 | ···················· | ···················· | ···················· | ···················· | ···················· | ···················· |
| N | 2805 | hCoV-19/USA/CO-QDX-2368/2020\|EPI_ISL_604688\|2020-0 | ···················· | ···················· | ···················· | ···················· | ···················· | ···················· |
| N | 2806 | hCoV-19/USA/CO-QDX-2378/2020\|EPI_ISL_604689\|2020-0 | ···················· | ···················· | ···················· | ···················· | ···················· | ···················· |
| N | 2807 | hCoV-19/USA/CO-QDX-2401/2020\|EPI_ISL_604690\|2020-0 | ···················· | ···················· | ···················· | ···················· | ···················· | ···················· |
| N | 2808 | hCoV-19/USA/CO-QDX-2403/2020\|EPI_ISL_604691\|2020-0 | ···················· | ···················· | ···················· | ···················· | ···················· | ···················· |
| N | 2809 | hCoV-19/USA/GA-QDX-2367/2020\|EPI_ISL_604692\|2020-0 | ···················· | ···················· | ···················· | ···················· | ···················· | ···················· |
| N | 2810 | hCoV-19/USA/GA-QDX-2379/2020\|EPI_ISL_604693\|2020-0 | ···················· | ···················· | ···················· | ···················· | ···················· | ···················· |
| N | 2811 | hCoV-19/USA/GA-QDX-2388/2020\|EPI_ISL_604694\|2020-0 | ···················· | ···················· | ···················· | ···················· | ···················· | ···················· |
| N | 2812 | hCoV-19/USA/GA-QDX-2420/2020\|EPI_ISL_604695\|2020-0 | ···················· | ···················· | ···················· | ···················· | ···················· | ···················· |
| N | 2813 | hCoV-19/USA/GA-QDX-2421/2020\|EPI_ISL_604696\|2020-0 | ···················· | ···················· | ···················· | ···················· | ···················· | ···················· |
| N | 2814 | hCoV-19/USA/GA-QDX-2425/2020\|EPI_ISL_604697\|2020-0 | ···················· | ···················· | ···················· | ···················· | ···················· | ···················· |
| N | 2815 | hCoV-19/USA/GA-QDX-2426/2020\|EPI_ISL_604698\|2020-0 | ···················· | ···················· | ···················· | ···················· | ···················· | ···················· |
| N | 2816 | hCoV-19/USA/MI-QDX-2316/2020\|EPI_ISL_604699\|2020-0 | ···················· | ···················· | ···················· | ···················· | ···················· | ···················· |
| N | 2817 | hCoV-19/England/CAMC-A583AC/2020\|EPI_ISL_597000\|20 | ···················· | ···················· | ···················· | ···················· | ···················· | ···················· |
| N | 2818 | hCoV-19/England/CAMC-A5950F/2020\|EPI_ISL_597001\|20 | ···················· | ···················· | ···················· | ···················· | ···················· | ···················· |
| N | 2819 | hCoV-19/England/CAMC-A58773/2020\|EPI_ISL_597002\|20 | ···················· | ···················· | ···················· | ···················· | ···················· | ···················· |
| N | 2820 | hCoV-19/England/CAMC-A5899B/2020\|EPI_ISL_597003\|20 | ···················· | ···················· | ···················· | ···················· | ···················· | ···················· |
| N | 2821 | hCoV-19/England/CAMC-A58746/2020\|EPI_ISL_597004\|20 | ···················· | ···················· | ···················· | ···················· | ···················· | ···················· |
| N | 2822 | hCoV-19/England/CAMC-A58F5D/2020\|EPI_ISL_597005\|20 | ···················· | ···················· | ···················· | ···················· | ···················· | ···················· |
| N | 2823 | hCoV-19/England/CAMC-A58E05/2020\|EPI_ISL_597006\|20 | ···················· | ···················· | ···················· | ···················· | ···················· | ···················· |
| N | 2824 | hCoV-19/England/CAMC-A58B0E/2020\|EPI_ISL_597008\|20 | ···················· | ···················· | ···················· | ···················· | ···················· | ···················· |
| N | 2825 | hCoV-19/England/CAMC-A59314/2020\|EPI_ISL_597009\|20 | ···················· | ···················· | ···················· | ···················· | ···················· | ···················· |
| N | 2826 | hCoV-19/England/CAMC-A5943F/2020\|EPI_ISL_597010\|20 | ···················· | ···················· | ···················· | ···················· | ···················· | ···················· |
| N | 2827 | hCoV-19/England/CAMC-A58D26/2020\|EPI_ISL_597011\|20 | ···················· | ···················· | ···················· | ···················· | ···················· | ···················· |
| N | 2828 | hCoV-19/England/CAMC-A57EE8/2020\|EPI_ISL_597012\|20 | ···················· | ···················· | ···················· | ···················· | ···················· | ···················· |
| N | 2829 | hCoV-19/England/CAMC-A57CEE/2020\|EPI_ISL_597013\|20 | ···················· | ···················· | ···················· | ···················· | ···················· | ···················· |
| N | 2830 | hCoV-19/England/CAMC-A58360/2020\|EPI_ISL_597014\|20 | ···················· | ···················· | ···················· | ···················· | ···················· | ···················· |
| N | 2831 | hCoV-19/England/CAMC-A58421/2020\|EPI_ISL_597015\|20 | ···················· | ···················· | ···················· | ···················· | ···················· | ···················· |
| N | 2832 | hCoV-19/England/CAMC-A58AD4/2020\|EPI_ISL_597016\|20 | ···················· | ···················· | ···················· | ···················· | ···················· | ···················· |
| N | 2833 | hCoV-19/England/CAMC-A58676/2020\|EPI_ISL_597018\|20 | ···················· | ···················· | ···················· | ···················· | ···················· | ···················· |
| N | 2834 | hCoV-19/England/CAMC-A58B59/2020\|EPI_ISL_597019\|20 | ···················· | ·A·················· | ···················· | ···················· | ···················· | ··A················· |
| N | 2835 | hCoV-19/England/CAMC-A5937E/2020\|EPI_ISL_597020\|20 | ···················· | ···················· | ···················· | ···················· | ···················· | ···················· |
| N | 2836 | hCoV-19/England/CAMC-A5951E/2020\|EPI_ISL_597021\|20 | ···················· | ···················· | ···················· | ···················· | ···················· | ···················· |
| N | 2837 | hCoV-19/England/CAMC-A57FE5/2020\|EPI_ISL_597022\|20 | ···················· | ···················· | ···················· | ···················· | ···················· | ···················· |
| N | 2838 | hCoV-19/England/CAMC-A580E1/2020\|EPI_ISL_597023\|20 | ···················· | ···················· | ···················· | ···················· | ···················· | ···················· |
| N | 2839 | hCoV-19/England/CAMC-A583D9/2020\|EPI_ISL_597024\|20 | ···················· | ···················· | ···················· | ···················· | ···················· | ···················· |
| N | 2840 | hCoV-19/England/CAMC-A58658/2020\|EPI_ISL_597025\|20 | ···················· | ···················· | ···················· | ···················· | ···················· | ···················· |
| N | 2841 | hCoV-19/England/CAMC-A596C0/2020\|EPI_ISL_597027\|20 | ···················· | ···················· | ···················· | ···················· | ·····K·············· | ···················· |
| N | 2842 | hCoV-19/England/CAMC-A57E51/2020\|EPI_ISL_597028\|20 | ···················· | ···················· | ···················· | ···················· | ···················· | ···················· |
| N | 2843 | hCoV-19/England/CAMC-A59077/2020\|EPI_ISL_597030\|20 | ···················· | ···················· | ···················· | ···················· | ···················· | ···················· |
| N | 2844 | hCoV-19/England/CAMC-A5805A/2020\|EPI_ISL_597031\|20 | ···················· | ···················· | ···················· | ···················· | ···················· | ···················· |
| N | 2845 | hCoV-19/England/CAMC-A58CFC/2020\|EPI_ISL_597032\|20 | ···················· | ···················· | ···················· | ···················· | ···T················ | ···················· |
| N | 2846 | hCoV-19/England/CAMC-A59736/2020\|EPI_ISL_597033\|20 | ···················· | ·A·················· | ···················· | ···················· | ···················· | ···················· |
| N | 2847 | hCoV-19/England/CAMC-A57FD6/2020\|EPI_ISL_597034\|20 | ···················· | ···················· | ···················· | ···················· | ···················· | ···················· |
| N | 2848 | hCoV-19/England/CAMC-A58861/2020\|EPI_ISL_597035\|20 | ···················· | ···················· | ···················· | ···················· | ···················· | ···················· |
| N | 2849 | hCoV-19/England/CAMC-A58597/2020\|EPI_ISL_597036\|20 | ···················· | ···················· | ···················· | ···················· | ···················· | ···················· |
| N | 2850 | hCoV-19/England/CAMC-A596FD/2020\|EPI_ISL_597037\|20 | ···················· | ···················· | ···················· | ···················· | ···················· | ···················· |
| N | 2851 | hCoV-19/England/ALDP-A518A4/2020\|EPI_ISL_598152\|20 | ···················· | ···················· | ···················· | ···················· | ···················· | ···················· |
| N | 2852 | hCoV-19/England/ALDP-A513C1/2020\|EPI_ISL_598153\|20 | ···················· | ···················· | ···················· | ···················· | ···················· | ···················· |
| N | 2853 | hCoV-19/England/MILK-A474F8/2020\|EPI_ISL_598300\|20 | ···················· | ···················· | ···················· | ···················· | ···················· | ···················· |
| N | 2854 | hCoV-19/England/MILK-A47D75/2020\|EPI_ISL_598301\|20 | ···················· | ···················· | ···················· | ···················· | ···················· | ···················· |
| N | 2855 | hCoV-19/England/MILK-A4721C/2020\|EPI_ISL_598302\|20 | ···················· | ···················· | ···················· | ···················· | ···················· | ···················· |
| N | 2856 | hCoV-19/England/MILK-A47461/2020\|EPI_ISL_598304\|20 | ···················· | ···················· | ···················· | ···················· | ···················· | ···················· |
| N | 2857 | hCoV-19/England/MILK-A4755F/2020\|EPI_ISL_598306\|20 | ···················· | ···················· | ···················· | ···················· | ·····T·············· | ···················· |
| N | 2858 | hCoV-19/England/MILK-A476B6/2020\|EPI_ISL_598307\|20 | ···················· | ···················· | ···················· | ···················· | ···················· | ···················· |
| N | 2859 | hCoV-19/England/MILK-A471D3/2020\|EPI_ISL_598308\|20 | ···················· | ···················· | ···················· | ···················· | ···················· | ···················· |
| N | 2860 | hCoV-19/England/MILK-A47A14/2020\|EPI_ISL_598314\|20 | ···················· | ···················· | ···················· | ···················· | ···················· | ···················· |
| N | 2861 | hCoV-19/England/MILK-A47ECD/2020\|EPI_ISL_598315\|20 | ···················· | ···················· | ···················· | ···················· | ···················· | ···················· |
| N | 2862 | hCoV-19/England/MILK-A46EA0/2020\|EPI_ISL_598316\|20 | ···················· | ···················· | ···················· | ···················· | ·····T·············· | ···················· |
| N | 2863 | hCoV-19/England/MILK-A4713D/2020\|EPI_ISL_598317\|20 | ···················· | ···················· | ···················· | ···················· | ···················· | ···················· |
| N | 2864 | hCoV-19/England/MILK-A47C5A/2020\|EPI_ISL_598318\|20 | ···················· | ·A·················· | ···················· | ···················· | ···················· | ··A················· |
| N | 2865 | hCoV-19/England/MILK-A47319/2020\|EPI_ISL_598319\|20 | ···················· | ···················· | ···················· | ···················· | ···················· | ···················· |
| N | 2866 | hCoV-19/England/MILK-A4705E/2020\|EPI_ISL_598320\|20 | ···················· | ···················· | ···················· | ···················· | ···················· | ···················· |
| N | 2867 | hCoV-19/England/MILK-A468EE/2020\|EPI_ISL_598321\|20 | ···················· | ···················· | ···················· | ···················· | ···················· | ···················· |

| | | | | | | | | |
|---|---|---|---|---|---|---|---|---|
| N | 2868 | hCoV-19/England/MILK-A479AE/2020\|EPI_ISL_598323\|20 | ···················· | ···················· | ···················· | ···················· | ······T············· | ···················· |
| N | 2869 | hCoV-19/England/MILK-A472A3/2020\|EPI_ISL_598324\|20 | ···················· | ···················· | ···················· | ···················· | ···················· | ···················· |
| N | 2870 | hCoV-19/Wales/MILK-A46C1F/2020\|EPI_ISL_598325\|2020 | ···················· | ···················· | ···················· | ···················· | ···················· | ···················· |
| N | 2871 | hCoV-19/England/MILK-A46C6A/2020\|EPI_ISL_598326\|20 | ···················· | ···················· | ···················· | ···················· | ···················· | ···················· |
| N | 2872 | hCoV-19/England/MILK-A469FA/2020\|EPI_ISL_598327\|20 | ···················· | ···················· | ···················· | ···················· | ···················· | ···················· |
| N | 2873 | hCoV-19/England/MILK-A46A06/2020\|EPI_ISL_598329\|20 | ···················· | ····G··············· | ···················· | ···················· | ···················· | ···················· |
| N | 2874 | hCoV-19/England/MILK-A46D76/2020\|EPI_ISL_598332\|20 | ···················· | ···················· | ···················· | ···················· | ···················· | ···················· |
| N | 2875 | hCoV-19/England/MILK-A46C4C/2020\|EPI_ISL_598333\|20 | ···················· | ·A·················· | ···················· | ···················· | ···················· | ···················· |
| N | 2876 | hCoV-19/England/MILK-A4715B/2020\|EPI_ISL_598334\|20 | ···················· | ···················· | ···················· | ···················· | ···················· | ···················· |
| N | 2877 | hCoV-19/England/MILK-A47276/2020\|EPI_ISL_598335\|20 | ···················· | ···················· | ···················· | ···················· | ···················· | ···················· |
| N | 2878 | hCoV-19/England/MILK-A4714C/2020\|EPI_ISL_598336\|20 | ···················· | ···················· | ···················· | ···················· | ···················· | ···················· |
| N | 2879 | hCoV-19/England/MILK-A47B20/2020\|EPI_ISL_598337\|20 | ···················· | ···················· | ···················· | ···················· | ···················· | ···················· |
| N | 2880 | hCoV-19/England/MILK-A46E73/2020\|EPI_ISL_598338\|20 | ···················· | ···················· | ···················· | ···················· | ···················· | ···················· |
| N | 2881 | hCoV-19/England/MILK-A475AA/2020\|EPI_ISL_598339\|20 | ···················· | ···················· | ···················· | ···················· | ······T············· | ···················· |
| N | 2882 | hCoV-19/England/MILK-A4806C/2020\|EPI_ISL_598340\|20 | ···················· | ···················· | ···················· | ···················· | ···················· | ···················· |
| N | 2883 | hCoV-19/England/MILK-A47416/2020\|EPI_ISL_598341\|20 | ···················· | ···················· | ···················· | ···················· | ···················· | ···················· |
| N | 2884 | hCoV-19/England/MILK-A47847/2020\|EPI_ISL_598343\|20 | ···················· | ·A·················· | ···················· | ···················· | ···················· | ···A················ |
| N | 2885 | hCoV-19/England/MILK-A4799F/2020\|EPI_ISL_598344\|20 | ···················· | ···················· | ···················· | ···················· | ···················· | ···················· |
| N | 2886 | hCoV-19/England/MILK-A475F5/2020\|EPI_ISL_598346\|20 | ···················· | ···················· | ···················· | ···················· | ···················· | ···················· |
| N | 2887 | hCoV-19/England/MILK-A47373/2020\|EPI_ISL_598347\|20 | ···················· | ···················· | ···················· | ···················· | ···················· | ···················· |
| N | 2888 | hCoV-19/England/MILK-A47FCA/2020\|EPI_ISL_598348\|20 | ···················· | ···················· | ···················· | ···················· | ···················· | ···················· |
| N | 2889 | hCoV-19/England/MILK-A47179/2020\|EPI_ISL_598349\|20 | ···················· | ···················· | ···················· | ···················· | ···················· | ···················· |
| N | 2890 | hCoV-19/England/MILK-A4708B/2020\|EPI_ISL_598351\|20 | ···················· | ···················· | ···················· | ···················· | ···················· | ···················· |
| N | 2891 | hCoV-19/England/MILK-A46963/2020\|EPI_ISL_598352\|20 | ···················· | ···················· | ···················· | ···················· | ···················· | ···················· |
| N | 2892 | hCoV-19/England/MILK-A47021/2020\|EPI_ISL_598353\|20 | ···················· | ···················· | ···················· | ···················· | ···················· | ···················· |
| N | 2893 | hCoV-19/England/MILK-A46B03/2020\|EPI_ISL_598354\|20 | ···················· | ···················· | ···················· | ···················· | ···················· | ···················· |
| N | 2894 | hCoV-19/England/MILK-A46A42/2020\|EPI_ISL_598355\|20 | ···················· | ···················· | ···················· | ···················· | ···················· | ···················· |
| N | 2895 | hCoV-19/England/MILK-A478B0/2020\|EPI_ISL_598356\|20 | ···················· | ···················· | ···················· | ···················· | ···················· | ···················· |
| N | 2896 | hCoV-19/England/MILK-A46AF7/2020\|EPI_ISL_598357\|20 | ···················· | ·A·················· | ···················· | ···················· | ···················· | ···················· |
| N | 2897 | hCoV-19/England/MILK-A478ED/2020\|EPI_ISL_598358\|20 | ···················· | ···················· | ···················· | ···················· | ······T············· | ···················· |
| N | 2898 | hCoV-19/England/MILK-A47980/2020\|EPI_ISL_598359\|20 | ···················· | ···················· | ···················· | ···················· | ···················· | ···················· |
| N | 2899 | hCoV-19/England/MILK-A46F43/2020\|EPI_ISL_598360\|20 | ···················· | ···················· | ···················· | ···················· | ···················· | ···················· |
| N | 2900 | hCoV-19/England/MILK-A47F8E/2020\|EPI_ISL_598362\|20 | ···················· | ···················· | ···················· | ···················· | ···················· | ···················· |
| N | 2901 | hCoV-19/England/MILK-A4804E/2020\|EPI_ISL_598363\|20 | ···················· | ···················· | ···················· | ···················· | ···················· | ···················· |
| N | 2902 | hCoV-19/England/MILK-A46F61/2020\|EPI_ISL_598364\|20 | ···················· | ···················· | ···················· | ···················· | ···················· | ···················· |
| N | 2903 | hCoV-19/England/MILK-A47768/2020\|EPI_ISL_598365\|20 | ···················· | ···················· | ···················· | ···················· | ···················· | ···················· |
| N | 2904 | hCoV-19/England/MILK-A47540/2020\|EPI_ISL_598366\|20 | ···················· | ···················· | ···················· | ···················· | ···················· | ···················· |
| N | 2905 | hCoV-19/England/QEUH-9DDEFA/2020\|EPI_ISL_599600\|20 | ···················· | ···················· | ···················· | ···················· | ····T··············· | ···················· |
| N | 2906 | hCoV-19/England/QEUH-9DDF06/2020\|EPI_ISL_599601\|20 | ···················· | ···················· | ···················· | ···················· | ······T············· | ···················· |
| N | 2907 | hCoV-19/England/QEUH-9DDE18/2020\|EPI_ISL_599602\|20 | ···················· | ···················· | ···················· | ···················· | ···················· | ···················· |
| N | 2908 | hCoV-19/England/QEUH-9DE4CA/2020\|EPI_ISL_599605\|20 | ···················· | ···················· | ···················· | ···················· | ···················· | ···················· |
| N | 2909 | hCoV-19/England/QEUH-9DDA9C/2020\|EPI_ISL_599606\|20 | ···················· | ···················· | ···················· | ···················· | ···················· | ···················· |
| N | 2910 | hCoV-19/Scotland/QEUH-9DDC0F/2020\|EPI_ISL_599607\|2 | ···················· | ···················· | ···················· | ···················· | ···················· | ···················· |
| N | 2911 | hCoV-19/England/QEUH-9DDE27/2020\|EPI_ISL_599608\|20 | ···················· | ·A·················· | ···················· | ···················· | ···················· | ···A················ |
| N | 2912 | hCoV-19/England/QEUH-9DDCA5/2020\|EPI_ISL_599610\|20 | ···················· | ···················· | ···················· | ···················· | ···················· | ···················· |
| N | 2913 | hCoV-19/England/QEUH-9DD67A/2020\|EPI_ISL_599611\|20 | ···················· | ···················· | ···················· | ···················· | ···················· | ···················· |
| N | 2914 | hCoV-19/England/QEUH-9DD99F/2020\|EPI_ISL_599612\|20 | ···················· | ·A·················· | ···················· | ···················· | ···················· | ···················· |
| N | 2915 | hCoV-19/England/QEUH-9DDFD9/2020\|EPI_ISL_599613\|20 | ···················· | ···················· | ···················· | ···················· | ···················· | ···················· |
| N | 2916 | hCoV-19/England/QEUH-9DE65B/2020\|EPI_ISL_599614\|20 | ···················· | ···················· | ···················· | ···················· | ···················· | ···················· |
| N | 2917 | hCoV-19/England/QEUH-9DD917/2020\|EPI_ISL_599615\|20 | ···················· | ···················· | ···················· | ···················· | ···················· | ···················· |
| N | 2918 | hCoV-19/England/QEUH-9DDF60/2020\|EPI_ISL_599617\|20 | ···················· | ···················· | ···················· | ···················· | ···················· | ···················· |
| N | 2919 | hCoV-19/England/QEUH-9DD865/2020\|EPI_ISL_599618\|20 | ···················· | ···················· | ···················· | ···················· | ····T··············· | ···················· |
| N | 2920 | hCoV-19/England/QEUH-9DD021/2020\|EPI_ISL_599619\|20 | ···················· | ···················· | ···················· | ···················· | ···················· | ···················· |
| N | 2921 | hCoV-19/England/QEUH-9DD434/2020\|EPI_ISL_599620\|20 | ···················· | ···················· | ···················· | ···················· | ···················· | ···················· |
| N | 2922 | hCoV-19/England/QEUH-9DDBB7/2020\|EPI_ISL_599621\|20 | ···················· | ···················· | ···················· | ···················· | ···················· | ···················· |
| N | 2923 | hCoV-19/England/QEUH-9DDB11/2020\|EPI_ISL_599622\|20 | ···················· | ···················· | ···················· | ···················· | ···················· | ···················· |
| N | 2924 | hCoV-19/England/QEUH-9DCCC4/2020\|EPI_ISL_599623\|20 | ···················· | ···················· | ···················· | ···················· | ···················· | ···················· |
| N | 2925 | hCoV-19/England/QEUH-9DDABA/2020\|EPI_ISL_599624\|20 | ···················· | ···················· | ···················· | ···················· | ···················· | ···················· |
| N | 2926 | hCoV-19/England/QEUH-9DE196/2020\|EPI_ISL_599625\|20 | ···················· | ···················· | ···················· | ···················· | ···················· | ···················· |
| N | 2927 | hCoV-19/England/QEUH-9DE390/2020\|EPI_ISL_599626\|20 | ···················· | ···················· | ···················· | ···················· | ···················· | ···················· |
| N | 2928 | hCoV-19/Scotland/QEUH-9DD65C/2020\|EPI_ISL_599629\|2 | ···················· | ···················· | ···················· | ···················· | ···················· | ···················· |
| N | 2929 | hCoV-19/England/QEUH-9DD610/2020\|EPI_ISL_599630\|20 | ···················· | ···················· | ···················· | ···················· | ···················· | ···················· |
| N | 2930 | hCoV-19/England/QEUH-9DDE09/2020\|EPI_ISL_599631\|20 | ···················· | ···················· | ···················· | ···················· | ···················· | ···················· |
| N | 2931 | hCoV-19/England/QEUH-9DDF33/2020\|EPI_ISL_599632\|20 | ···················· | ···················· | ···················· | ···················· | ···················· | ···················· |
| N | 2932 | hCoV-19/England/QEUH-9DDF7F/2020\|EPI_ISL_599633\|20 | ···················· | ···················· | ···················· | ···················· | ···················· | ···················· |
| N | 2933 | hCoV-19/England/QEUH-9DE14B/2020\|EPI_ISL_599634\|20 | ···················· | ···················· | ···················· | ···················· | ···················· | ···················· |
| N | 2934 | hCoV-19/Scotland/QEUH-9DCC97/2020\|EPI_ISL_599635\|2 | ···················· | ···················· | ···················· | ···················· | ···················· | ···················· |
| N | 2935 | hCoV-19/England/QEUH-9DE3DC/2020\|EPI_ISL_599636\|20 | ···················· | ···················· | ···················· | ···················· | ···················· | ···················· |
| N | 2936 | hCoV-19/England/QEUH-9DE372/2020\|EPI_ISL_599637\|20 | ···················· | ···················· | ···················· | ···················· | ···················· | ···················· |
| N | 2937 | hCoV-19/England/QEUH-9DDE36/2020\|EPI_ISL_599638\|20 | ···················· | ·A·················· | ···················· | ···················· | ···················· | ···A················ |
| N | 2938 | hCoV-19/England/QEUH-9DD6A7/2020\|EPI_ISL_599639\|20 | ···················· | ···················· | ···················· | ···················· | ···················· | ···················· |
| N | 2939 | hCoV-19/England/QEUH-9DD5C8/2020\|EPI_ISL_599645\|20 | ···················· | ···················· | ···················· | ···················· | ····T··············· | ···················· |
| N | 2940 | hCoV-19/Scotland/QEUH-9DD8A1/2020\|EPI_ISL_599646\|2 | ···················· | ···················· | ···················· | ···················· | ···················· | ···················· |
| N | 2941 | hCoV-19/Scotland/QEUH-9DD16A/2020\|EPI_ISL_599647\|2 | ···················· | ···················· | ···················· | ···················· | ···················· | ···················· |
| N | 2942 | hCoV-19/England/QEUH-9DD003/2020\|EPI_ISL_599649\|20 | ···················· | ···················· | ···················· | ···················· | ···················· | ···················· |
| N | 2943 | hCoV-19/England/QEUH-9DDFF7/2020\|EPI_ISL_599650\|20 | ···················· | ···················· | ···················· | ···················· | ···················· | ···················· |
| N | 2944 | hCoV-19/England/QEUH-9DCF70/2020\|EPI_ISL_599651\|20 | ···················· | ···················· | ···················· | ···················· | ···················· | ···················· |
| N | 2945 | hCoV-19/England/QEUH-9DDB7B/2020\|EPI_ISL_599652\|20 | ···················· | ···················· | ···················· | ···················· | ···················· | ···················· |
| N | 2946 | hCoV-19/England/QEUH-9DCF43/2020\|EPI_ISL_599653\|20 | ···················· | ·A·················· | ···················· | ···················· | ···················· | ···A················ |
| N | 2947 | hCoV-19/England/QEUH-9DDAC9/2020\|EPI_ISL_599654\|20 | ···················· | ···················· | ···················· | ···················· | ···················· | ···················· |

| | | | | | | | | |
|---|---|---|---|---|---|---|---|---|
| N | 2948 | hCoV-19/England/QEUH-9DE442/2020\|EPI_ISL_599655\|20 | ···················· | ·A·················· | ····················· | ···················· | ···················· | ··A················· |
| N | 2949 | hCoV-19/England/QEUH-9DDC1E/2020\|EPI_ISL_599656\|20 | ···················· | ···················· | ····················· | ···················· | ···················· | ···················· |
| N | 2950 | hCoV-19/England/QEUH-9DD08B/2020\|EPI_ISL_599657\|20 | ···················· | ···················· | ····················· | ···················· | ···················· | ···················· |
| N | 2951 | hCoV-19/Scotland/QEUH-9DD504/2020\|EPI_ISL_599658\|2 | ···················· | ···················· | ····················· | ···················· | ···················· | ···················· |
| N | 2952 | hCoV-19/England/QEUH-9DCCD3/2020\|EPI_ISL_599659\|20 | ···················· | ···················· | ····················· | ···················· | ···················· | ···················· |
| N | 2953 | hCoV-19/England/QEUH-9DD64D/2020\|EPI_ISL_599660\|20 | ···················· | ···················· | ····················· | ···················· | ···················· | ···················· |
| N | 2954 | hCoV-19/England/QEUH-9DD9CC/2020\|EPI_ISL_599661\|20 | ···················· | ···················· | ····················· | ···················· | ···················· | ···················· |
| N | 2955 | hCoV-19/England/QEUH-9DE55E/2020\|EPI_ISL_599662\|20 | ···················· | ···················· | ····················· | ···················· | ···················· | ···················· |
| N | 2956 | hCoV-19/England/QEUH-9DCF9E/2020\|EPI_ISL_599664\|20 | ···················· | ···················· | ····················· | ···················· | ···················· | ···················· |
| N | 2957 | hCoV-19/England/QEUH-9DDA50/2020\|EPI_ISL_599665\|20 | ···················· | ···················· | ····················· | ···················· | ···T················ | ···················· |
| N | 2958 | hCoV-19/England/QEUH-9DDAAB/2020\|EPI_ISL_599666\|20 | ···················· | ···················· | ····················· | ···················· | ···T················ | ···················· |
| N | 2959 | hCoV-19/England/QEUH-9DE293/2020\|EPI_ISL_599668\|20 | ···················· | ·A·················· | ····················· | ···················· | ···················· | ··A················· |
| N | 2960 | hCoV-19/England/QEUH-9DCF34/2020\|EPI_ISL_599669\|20 | ···················· | ···················· | ····················· | ···················· | ···················· | ···················· |
| N | 2961 | hCoV-19/England/QEUH-9DE099/2020\|EPI_ISL_599670\|20 | ···················· | ···················· | ····················· | ···················· | ···················· | ···················· |
| N | 2962 | hCoV-19/Scotland/QEUH-9DD407/2020\|EPI_ISL_599671\|2 | ···················· | ···················· | ····················· | ···················· | ···················· | ···················· |
| N | 2963 | hCoV-19/England/QEUH-9DDD93/2020\|EPI_ISL_599672\|20 | ···················· | ···················· | ····················· | ···················· | ···················· | ···················· |
| N | 2964 | hCoV-19/England/QEUH-9DDF51/2020\|EPI_ISL_599673\|20 | ···················· | ·A·················· | ····················· | ···················· | ···················· | ···················· |
| N | 2965 | hCoV-19/England/QEUH-9DE06C/2020\|EPI_ISL_599674\|20 | ···················· | ·A·················· | ····················· | ···················· | ···················· | ··A················· |
| N | 2966 | hCoV-19/England/QEUH-9DD838/2020\|EPI_ISL_599675\|20 | ···················· | ···················· | ····················· | ···················· | ···T················ | ···················· |
| N | 2967 | hCoV-19/Scotland/QEUH-9DD522/2020\|EPI_ISL_599676\|2 | ···················· | ···················· | ····················· | ···················· | ···················· | ···················· |
| N | 2968 | hCoV-19/Scotland/QEUH-9DD15B/2020\|EPI_ISL_599677\|2 | ···················· | ···················· | ····················· | ···················· | ···················· | ···················· |
| N | 2969 | hCoV-19/England/QEUH-9DCE55/2020\|EPI_ISL_599678\|20 | ···················· | ···················· | ····················· | ···················· | ···················· | ···················· |
| N | 2970 | hCoV-19/England/QEUH-9DD58C/2020\|EPI_ISL_599679\|20 | ···················· | ···················· | ····················· | ···················· | ···················· | ···················· |
| N | 2971 | hCoV-19/England/QEUH-9DDC87/2020\|EPI_ISL_599680\|20 | ···················· | ···················· | ····················· | ···················· | ···················· | ···················· |
| N | 2972 | hCoV-19/England/QEUH-9DD1C4/2020\|EPI_ISL_599682\|20 | ···················· | ···················· | ····················· | ···················· | ···················· | ···················· |
| N | 2973 | hCoV-19/Scotland/QEUH-9DD4DA/2020\|EPI_ISL_599683\|2 | ···················· | ···················· | ····················· | ···················· | ···················· | ···················· |
| N | 2974 | hCoV-19/England/QEUH-9DCCF1/2020\|EPI_ISL_599684\|20 | ···················· | ···················· | ····················· | ···················· | ···················· | ···················· |
| N | 2975 | hCoV-19/Scotland/QEUH-9DD4F8/2020\|EPI_ISL_599685\|2 | ···················· | ···················· | ····················· | ···················· | ···················· | ···················· |
| N | 2976 | hCoV-19/England/QEUH-9DE257/2020\|EPI_ISL_599686\|20 | ···················· | ···················· | ····················· | ···················· | ···················· | ···················· |
| N | 2977 | hCoV-19/England/QEUH-9DDFE8/2020\|EPI_ISL_599687\|20 | ···················· | ···················· | ····················· | ···················· | ···················· | ···················· |
| N | 2978 | hCoV-19/England/QEUH-9DD935/2020\|EPI_ISL_599688\|20 | ···················· | ···················· | ····················· | ···················· | ···················· | ···················· |
| N | 2979 | hCoV-19/England/QEUH-9DDD0C/2020\|EPI_ISL_599689\|20 | ···················· | ···················· | ····················· | ···················· | ···················· | ···················· |
| N | 2980 | hCoV-19/England/QEUH-9DE424/2020\|EPI_ISL_599690\|20 | ···················· | ·A·················· | ····················· | ···················· | ···················· | ··A················· |
| N | 2981 | hCoV-19/Scotland/QEUH-9DD7C2/2020\|EPI_ISL_599691\|2 | ···················· | ···················· | ····················· | ···················· | ···················· | ···················· |
| N | 2982 | hCoV-19/England/QEUH-9DDB99/2020\|EPI_ISL_599692\|20 | ···················· | ·A·················· | ····················· | ···················· | ···················· | ··A················· |
| N | 2983 | hCoV-19/England/QEUH-9DDA41/2020\|EPI_ISL_599693\|20 | ···················· | ···················· | ····················· | ···················· | ···················· | ···················· |
| N | 2984 | hCoV-19/England/QEUH-9DD601/2020\|EPI_ISL_599694\|20 | ···················· | ···················· | ····················· | ···················· | ···················· | ···················· |
| N | 2985 | hCoV-19/England/QEUH-9DE54F/2020\|EPI_ISL_599695\|20 | ···················· | ···················· | ····················· | ···················· | ···················· | ···················· |
| N | 2986 | hCoV-19/England/QEUH-9DDD75/2020\|EPI_ISL_599696\|20 | ···················· | ···················· | ····················· | ···················· | ···················· | ···················· |
| N | 2987 | hCoV-19/Scotland/QEUH-9DD48F/2020\|EPI_ISL_599697\|2 | ···················· | ···················· | ····················· | ···················· | ·····T·············· | ···················· |
| N | 2988 | hCoV-19/Scotland/QEUH-9DD1B5/2020\|EPI_ISL_599698\|2 | ···················· | ···················· | ····················· | ···················· | ···················· | ···················· |
| N | 2989 | hCoV-19/England/204201048/2020\|EPI_ISL_603200\|2020 | ···················· | ···················· | ····················· | ···················· | ···················· | ···················· |
| N | 2990 | hCoV-19/England/204201049/2020\|EPI_ISL_603201\|2020 | ···················· | ···················· | ····················· | ···················· | ···················· | ···················· |
| N | 2991 | hCoV-19/England/204201057/2020\|EPI_ISL_603202\|2020 | ···················· | ···················· | ····················· | ···················· | ···················· | ···················· |
| N | 2992 | hCoV-19/England/204201059/2020\|EPI_ISL_603203\|2020 | ···················· | ···················· | ····················· | ···················· | ···················· | ···················· |
| N | 2993 | hCoV-19/England/204220520/2020\|EPI_ISL_603204\|2020 | ···················· | ···················· | ····················· | ···················· | ···················· | ···················· |
| N | 2994 | hCoV-19/England/204220524/2020\|EPI_ISL_603205\|2020 | ···················· | ···················· | ····················· | ···················· | ···················· | ···················· |
| N | 2995 | hCoV-19/England/204221297/2020\|EPI_ISL_603206\|2020 | ···················· | ···················· | ····················· | ···················· | ···················· | ···················· |
| N | 2996 | hCoV-19/England/204262881/2020\|EPI_ISL_603207\|2020 | ···················· | ···················· | ····················· | ···················· | ···················· | ···················· |
| N | 2997 | hCoV-19/England/204262884/2020\|EPI_ISL_603208\|2020 | ···················· | ···················· | ····················· | ···················· | ···················· | ···················· |
| N | 2998 | hCoV-19/England/204262885/2020\|EPI_ISL_603209\|2020 | ···················· | ···················· | ····················· | ···················· | ···················· | ···················· |
| N | 2999 | hCoV-19/England/204262886/2020\|EPI_ISL_603210\|2020 | ···················· | ···················· | ····················· | ···················· | ···················· | ···················· |
| N | 3000 | hCoV-19/England/204262887/2020\|EPI_ISL_603211\|2020 | ···················· | ···················· | ····················· | ···················· | ···················· | ···················· |
| N | 3001 | hCoV-19/England/204262899/2020\|EPI_ISL_603212\|2020 | ···················· | ···················· | ····················· | ···················· | ···················· | ···················· |
| N | 3002 | hCoV-19/England/204262900/2020\|EPI_ISL_603213\|2020 | ···················· | ···················· | ····················· | ···················· | ···················· | ···················· |
| N | 3003 | hCoV-19/England/204262903/2020\|EPI_ISL_603214\|2020 | ···················· | ···················· | ····················· | ···················· | ···················· | ···················· |
| N | 3004 | hCoV-19/France/OCC-76/2020\|EPI_ISL_603216\|2020-09- | ···················· | ···················· | ····················· | ···················· | ···················· | ···················· |
| N | 3005 | hCoV-19/France/OCC-77/2020\|EPI_ISL_603217\|2020-09- | ···················· | ···················· | ····················· | ···················· | ···················· | ···················· |
| N | 3006 | hCoV-19/France/OCC-82/2020\|EPI_ISL_603218\|2020-09- | ···················· | ···················· | ····················· | ···················· | ···················· | ···················· |
| N | 3007 | hCoV-19/France/OCC-83/2020\|EPI_ISL_603219\|2020-09- | ···················· | ···················· | ····················· | ···················· | ···················· | ···················· |
| N | 3008 | hCoV-19/France/OCC-84/2020\|EPI_ISL_603220\|2020-09- | ···················· | ···················· | ····················· | ···················· | ···················· | ···················· |
| N | 3009 | hCoV-19/Bangladesh/BCSIR-NILMRC_392/2020\|EPI_ISL_6 | ···················· | ···················· | ····················· | ···················· | ···················· | ···················· |
| N | 3010 | hCoV-19/Bangladesh/BCSIR-NILMRC_398/2020\|EPI_ISL_6 | ···················· | ···················· | ····················· | ···················· | ···················· | ···················· |
| N | 3011 | hCoV-19/Bangladesh/BCSIR-NILMRC_376A/2020\|EPI_ISL_ | ···················· | ···················· | ····················· | ···················· | ···················· | ···················· |
| N | 3012 | hCoV-19/Bangladesh/BCSIR-NILMRC_390/2020\|EPI_ISL_6 | ···················· | ···················· | ····················· | ···················· | ···················· | ··A················· |
| N | 3013 | hCoV-19/Bangladesh/BCSIR-NILMRC_391/2020\|EPI_ISL_6 | ···················· | ···················· | ····················· | ···················· | ···················· | ···················· |
| N | 3014 | hCoV-19/USA/WI-QDX-2121/2020\|EPI_ISL_604500\|2020-0 | ···················· | ···················· | ····················· | ···················· | ···················· | ···················· |
| N | 3015 | hCoV-19/USA/AZ-QDX-2260/2020\|EPI_ISL_604501\|2020-0 | ···················· | ···················· | ····················· | ···················· | ···················· | ···················· |
| N | 3016 | hCoV-19/USA/AZ-QDX-2267/2020\|EPI_ISL_604502\|2020-0 | ···················· | ···················· | ····················· | ···················· | ···················· | ···················· |
| N | 3017 | hCoV-19/USA/AZ-QDX-2310/2020\|EPI_ISL_604503\|2020-0 | ···················· | ···················· | ····················· | ···················· | ···················· | ···················· |
| N | 3018 | hCoV-19/USA/AZ-QDX-2311/2020\|EPI_ISL_604504\|2020-0 | ···················· | ···················· | ····················· | ···················· | ···················· | ···················· |
| N | 3019 | hCoV-19/USA/CA-QDX-2203/2020\|EPI_ISL_604505\|2020-0 | ···················· | ···················· | ····················· | ···················· | ···················· | ···················· |
| N | 3020 | hCoV-19/USA/CA-QDX-2204/2020\|EPI_ISL_604506\|2020-0 | ···················· | ···················· | ····················· | ···················· | ···················· | ···················· |
| N | 3021 | hCoV-19/USA/CA-QDX-2226/2020\|EPI_ISL_604507\|2020-0 | ···················· | ···················· | ····················· | ···················· | ···················· | ···················· |
| N | 3022 | hCoV-19/USA/CA-QDX-2229/2020\|EPI_ISL_604508\|2020-0 | ···················· | ···················· | ····················· | ···················· | ···················· | ···················· |
| N | 3023 | hCoV-19/USA/CA-QDX-2244/2020\|EPI_ISL_604509\|2020-0 | ···················· | ···················· | ····················· | ···················· | ···················· | ···················· |
| N | 3024 | hCoV-19/USA/CA-QDX-2245/2020\|EPI_ISL_604510\|2020-0 | ···················· | ···················· | ····················· | ···················· | ···················· | ···················· |
| N | 3025 | hCoV-19/USA/CA-QDX-2256/2020\|EPI_ISL_604511\|2020-0 | ···················· | ···················· | ····················· | ···················· | ···················· | ···················· |
| N | 3026 | hCoV-19/USA/CA-QDX-2263/2020\|EPI_ISL_604512\|2020-0 | ···················· | ···················· | ····················· | ···················· | ···················· | ···················· |
| N | 3027 | hCoV-19/USA/CA-QDX-2275/2020\|EPI_ISL_604513\|2020-0 | ···················· | ···················· | ····················· | ···················· | ···················· | ···················· |

| | | | | | | | | |
|---|---|---|---|---|---|---|---|---|
| N | 3028 | hCoV-19/USA/CA-QDX-2282/2020\|EPI_ISL_604514\|2020-0 | ···················· | ···················· | ····················· | ···················· | ···················· | ···················· |
| N | 3029 | hCoV-19/USA/CA-QDX-2303/2020\|EPI_ISL_604515\|2020-0 | ···················· | ···················· | ····················· | ···················· | ···················· | ···················· |
| N | 3030 | hCoV-19/USA/CO-QDX-2195/2020\|EPI_ISL_604516\|2020-0 | ···················· | ···················· | ····················· | ···················· | ···················· | ···················· |
| N | 3031 | hCoV-19/USA/CO-QDX-2205/2020\|EPI_ISL_604517\|2020-0 | ···················· | ···················· | ····················· | ···················· | ···················· | ···················· |
| N | 3032 | hCoV-19/USA/CO-QDX-2225/2020\|EPI_ISL_604518\|2020-0 | ···················· | ···················· | ····················· | ···················· | ···················· | ···················· |
| N | 3033 | hCoV-19/USA/CO-QDX-2236/2020\|EPI_ISL_604519\|2020-0 | ···················· | ···················· | ····················· | ···················· | ···················· | ···················· |
| N | 3034 | hCoV-19/USA/CO-QDX-2254/2020\|EPI_ISL_604520\|2020-0 | ···················· | ···················· | ····················· | ···················· | ···················· | ···················· |
| N | 3035 | hCoV-19/USA/CO-QDX-2272/2020\|EPI_ISL_604521\|2020-0 | ···················· | ···················· | ····················· | ···················· | ···················· | ···················· |
| N | 3036 | hCoV-19/USA/CO-QDX-2274/2020\|EPI_ISL_604522\|2020-0 | ···················· | ···················· | ····················· | ···················· | ···················· | ···················· |
| N | 3037 | hCoV-19/USA/CO-QDX-2309/2020\|EPI_ISL_604523\|2020-0 | ···················· | ···················· | ····················· | ···················· | ···················· | ···················· |
| N | 3038 | hCoV-19/USA/CT-QDX-2200/2020\|EPI_ISL_604524\|2020-0 | ···················· | ···················· | ····················· | ···················· | ···················· | ···················· |
| N | 3039 | hCoV-19/USA/CT-QDX-2239/2020\|EPI_ISL_604525\|2020-0 | ···················· | ···················· | ····················· | ···················· | ···················· | ···················· |
| N | 3040 | hCoV-19/USA/CT-QDX-2279/2020\|EPI_ISL_604526\|2020-0 | ···················· | ···················· | ····················· | ···················· | ···················· | ···················· |
| N | 3041 | hCoV-19/USA/GA-QDX-2196/2020\|EPI_ISL_604527\|2020-0 | ···················· | ···················· | ····················· | ···················· | ···················· | ···················· |
| N | 3042 | hCoV-19/USA/GA-QDX-2209/2020\|EPI_ISL_604528\|2020-0 | ···················· | ···················· | ····················· | ···················· | ···················· | ···················· |
| N | 3043 | hCoV-19/USA/GA-QDX-2219/2020\|EPI_ISL_604529\|2020-0 | ···················· | ···················· | ····················· | ···················· | ···················· | ···················· |
| N | 3044 | hCoV-19/USA/GA-QDX-2235/2020\|EPI_ISL_604530\|2020-0 | ···················· | ···················· | ····················· | ···················· | ···················· | ···················· |
| N | 3045 | hCoV-19/USA/GA-QDX-2259/2020\|EPI_ISL_604531\|2020-0 | ···················· | ···················· | ····················· | ···················· | ···················· | ···················· |
| N | 3046 | hCoV-19/USA/IN-QDX-2220/2020\|EPI_ISL_604532\|2020-0 | ···················· | ···················· | ····················· | ···················· | ···················· | ···················· |
| N | 3047 | hCoV-19/USA/MD-QDX-2299/2020\|EPI_ISL_604534\|2020-0 | ···················· | ···················· | ····················· | ···················· | ···················· | ···················· |
| N | 3048 | hCoV-19/USA/MI-QDX-2193/2020\|EPI_ISL_604535\|2020-0 | ···················· | ···················· | ····················· | ···················· | ···················· | ···················· |
| N | 3049 | hCoV-19/USA/MI-QDX-2197/2020\|EPI_ISL_604536\|2020-0 | ···················· | ···················· | ····················· | ···················· | ···················· | ···················· |
| N | 3050 | hCoV-19/USA/MI-QDX-2199/2020\|EPI_ISL_604537\|2020-0 | ···················· | ···················· | ····················· | ···················· | ···················· | ···················· |
| N | 3051 | hCoV-19/USA/MI-QDX-2206/2020\|EPI_ISL_604538\|2020-0 | ···················· | ···················· | ····················· | ···················· | ···················· | ···················· |
| N | 3052 | hCoV-19/USA/MI-QDX-2212/2020\|EPI_ISL_604539\|2020-0 | ···················· | ···················· | ····················· | ···················· | ···················· | ···················· |
| N | 3053 | hCoV-19/USA/MI-QDX-2221/2020\|EPI_ISL_604540\|2020-0 | ···················· | ···················· | ····················· | ···················· | ···················· | ···················· |
| N | 3054 | hCoV-19/USA/MI-QDX-2232/2020\|EPI_ISL_604541\|2020-0 | ···················· | ···················· | ····················· | ···················· | ···················· | ···················· |
| N | 3055 | hCoV-19/USA/MI-QDX-2251/2020\|EPI_ISL_604542\|2020-0 | ···················· | ···················· | ····················· | ···················· | ···················· | ···················· |
| N | 3056 | hCoV-19/USA/MI-QDX-2252/2020\|EPI_ISL_604543\|2020-0 | ···················· | ···················· | ····················· | ···················· | ···················· | ···················· |
| N | 3057 | hCoV-19/USA/MI-QDX-2255/2020\|EPI_ISL_604544\|2020-0 | ···················· | ···················· | ····················· | ···················· | ···················· | ···················· |
| N | 3058 | hCoV-19/USA/MI-QDX-2261/2020\|EPI_ISL_604545\|2020-0 | ···················· | ···················· | ····················· | ···················· | ···················· | ···················· |
| N | 3059 | hCoV-19/USA/MI-QDX-2271/2020\|EPI_ISL_604546\|2020-0 | ···················· | ···················· | ····················· | ···················· | ···················· | ···················· |
| N | 3060 | hCoV-19/USA/MI-QDX-2284/2020\|EPI_ISL_604547\|2020-0 | ···················· | ···················· | ····················· | ···················· | ···················· | ···················· |
| N | 3061 | hCoV-19/USA/MI-QDX-2298/2020\|EPI_ISL_604548\|2020-0 | ···················· | ···················· | ····················· | ···················· | ···················· | ···················· |
| N | 3062 | hCoV-19/USA/MI-QDX-2306/2020\|EPI_ISL_604549\|2020-0 | ···················· | ···················· | ····················· | ···················· | ···················· | ···················· |
| N | 3063 | hCoV-19/USA/MI-QDX-2312/2020\|EPI_ISL_604550\|2020-0 | ···················· | ···················· | ····················· | ···················· | ···················· | ···················· |
| N | 3064 | hCoV-19/USA/MN-QDX-2258/2020\|EPI_ISL_604551\|2020-0 | ···················· | ···················· | ····················· | ···················· | ···················· | ···················· |
| N | 3065 | hCoV-19/USA/MN-QDX-2265/2020\|EPI_ISL_604552\|2020-0 | ···················· | ···················· | ····················· | ···················· | ···················· | ···················· |
| N | 3066 | hCoV-19/USA/NJ-QDX-2190/2020\|EPI_ISL_604553\|2020-0 | ···················· | ···················· | ····················· | ···················· | ···················· | ···················· |
| N | 3067 | hCoV-19/USA/NJ-QDX-2191/2020\|EPI_ISL_604554\|2020-0 | ···················· | ···················· | ····················· | ···················· | ···················· | ···················· |
| N | 3068 | hCoV-19/USA/NJ-QDX-2210/2020\|EPI_ISL_604555\|2020-0 | ···················· | ···················· | ····················· | ···················· | ···················· | ···················· |
| N | 3069 | hCoV-19/USA/NJ-QDX-2216/2020\|EPI_ISL_604556\|2020-0 | ···················· | ···················· | ····················· | ···················· | ···················· | ···················· |
| N | 3070 | hCoV-19/USA/NJ-QDX-2250/2020\|EPI_ISL_604557\|2020-0 | ···················· | ···················· | ····················· | ···················· | ···················· | ···················· |
| N | 3071 | hCoV-19/USA/NJ-QDX-2268/2020\|EPI_ISL_604558\|2020-0 | ···················· | ···················· | ····················· | ···················· | ···················· | ···················· |
| N | 3072 | hCoV-19/USA/NJ-QDX-2269/2020\|EPI_ISL_604559\|2020-0 | ···················· | ···················· | ····················· | ···················· | ···················· | ···················· |
| N | 3073 | hCoV-19/England/ALDP-A51121/2020\|EPI_ISL_598127\|20 | ···················· | ···················· | ····················· | ···················· | ···················· | ···················· |
| N | 3074 | hCoV-19/England/ALDP-A516D7/2020\|EPI_ISL_598128\|20 | ···················· | ···················· | ····················· | ···················· | ···················· | ···················· |
| N | 3075 | hCoV-19/England/ALDP-A5218A/2020\|EPI_ISL_598129\|20 | ···················· | ···················· | ····················· | ···················· | ···················· | ···················· |
| N | 3076 | hCoV-19/England/ALDP-A51C99/2020\|EPI_ISL_598130\|20 | ···················· | ···················· | ····················· | ···················· | ···················· | ···················· |
| N | 3077 | hCoV-19/England/ALDP-A52023/2020\|EPI_ISL_598131\|20 | ···················· | ···················· | ····················· | ···················· | ···················· | ···················· |
| N | 3078 | hCoV-19/England/ALDP-A524CD/2020\|EPI_ISL_598132\|20 | ···················· | ···················· | ····················· | ···················· | ···················· | ···················· |
| N | 3079 | hCoV-19/England/ALDP-A521B7/2020\|EPI_ISL_598134\|20 | ···················· | ···················· | ····················· | ···················· | ···················· | ···················· |
| N | 3080 | hCoV-19/England/ALDP-A5264F/2020\|EPI_ISL_598135\|20 | ···················· | ···················· | ····················· | ···················· | ···················· | ···················· |
| N | 3081 | hCoV-19/England/ALDP-A51B8D/2020\|EPI_ISL_598136\|20 | ···················· | ···················· | ····················· | ···················· | ···················· | ···················· |
| N | 3082 | hCoV-19/England/ALDP-A523C0/2020\|EPI_ISL_598137\|20 | ···················· | ···················· | ····················· | ···················· | ···················· | ···················· |
| N | 3083 | hCoV-19/England/ALDP-A51F09/2020\|EPI_ISL_598138\|20 | ···················· | ···················· | ····················· | ···················· | ···················· | ···················· |
| N | 3084 | hCoV-19/England/ALDP-A51D87/2020\|EPI_ISL_598139\|20 | ···················· | ···················· | ····················· | ···················· | ···················· | ···················· |
| N | 3085 | hCoV-19/England/ALDP-A5184A/2020\|EPI_ISL_598141\|20 | ···················· | ···················· | ····················· | ···················· | ···················· | ···················· |
| N | 3086 | hCoV-19/England/ALDP-A51983/2020\|EPI_ISL_598142\|20 | ···················· | ···················· | ····················· | ···················· | ···················· | ···················· |
| N | 3087 | hCoV-19/England/ALDP-A526B8/2020\|EPI_ISL_598143\|20 | ···················· | ···················· | ····················· | ···················· | ···················· | ···················· |
| N | 3088 | hCoV-19/England/ALDP-A527A6/2020\|EPI_ISL_598144\|20 | ···················· | ···················· | ····················· | ···················· | ···················· | ···················· |
| N | 3089 | hCoV-19/England/ALDP-A51DD2/2020\|EPI_ISL_598146\|20 | ···················· | ···················· | ····················· | ···················· | ···················· | ···················· |
| N | 3090 | hCoV-19/England/ALDP-A51455/2020\|EPI_ISL_598147\|20 | ···················· | ···················· | ····················· | ···················· | ···················· | ···················· |
| N | 3091 | hCoV-19/England/ALDP-A52384/2020\|EPI_ISL_598149\|20 | ···················· | ···················· | ····················· | ···················· | ···················· | ···················· |
| N | 3092 | hCoV-19/England/ALDP-A5217B/2020\|EPI_ISL_598150\|20 | ···················· | ···················· | ····················· | ···················· | ···················· | ···················· |
| N | 3093 | hCoV-19/England/ALDP-A52393/2020\|EPI_ISL_598151\|20 | ···················· | ···················· | ····················· | ···················· | ···················· | ···················· |
| N | 3094 | hCoV-19/England/ALDP-A5258E/2020\|EPI_ISL_598154\|20 | ···················· | ···················· | ····················· | ···················· | ···················· | ···················· |
| N | 3095 | hCoV-19/England/ALDP-A5140A/2020\|EPI_ISL_598155\|20 | ···················· | ···················· | ····················· | ···················· | ···················· | ···················· |
| N | 3096 | hCoV-19/England/ALDP-A51FFA/2020\|EPI_ISL_598156\|20 | ···················· | ···················· | ····················· | ···················· | ···················· | ···················· |
| N | 3097 | hCoV-19/England/ALDP-A5119A/2020\|EPI_ISL_598157\|20 | ···················· | ···················· | ····················· | ···················· | ···················· | ···················· |
| N | 3098 | hCoV-19/England/ALDP-A51F81/2020\|EPI_ISL_598159\|20 | ···················· | ···················· | ····················· | ···················· | ···················· | ···················· |
| N | 3099 | hCoV-19/England/ALDP-A51EEE/2020\|EPI_ISL_598160\|20 | ···················· | ···················· | ····················· | ···················· | ···················· | ···················· |
| N | 3100 | hCoV-19/England/ALDP-A5266D/2020\|EPI_ISL_598161\|20 | ···················· | ·A·················· | ····················· | ···················· | ···················· | ··A················· |
| N | 3101 | hCoV-19/England/ALDP-A5224B/2020\|EPI_ISL_598162\|20 | ···················· | ···················· | ····················· | ···················· | ···················· | ···················· |
| N | 3102 | hCoV-19/England/ALDP-A512E2/2020\|EPI_ISL_598164\|20 | ···················· | ···················· | ····················· | ···················· | ···················· | ···················· |
| N | 3103 | hCoV-19/England/ALDP-A51BBA/2020\|EPI_ISL_598165\|20 | ···················· | ···················· | ····················· | ···················· | ···················· | ···················· |
| N | 3104 | hCoV-19/England/ALDP-A5213F/2020\|EPI_ISL_598166\|20 | ···················· | ···················· | ····················· | ···················· | ···················· | ···················· |
| N | 3105 | hCoV-19/England/ALDP-A527E2/2020\|EPI_ISL_598168\|20 | ···················· | ···················· | ····················· | ···················· | ···················· | ···················· |
| N | 3106 | hCoV-19/England/ALDP-A515BC/2020\|EPI_ISL_598169\|20 | ···················· | ···················· | ····················· | ···················· | ···················· | ···················· |
| N | 3107 | hCoV-19/England/ALDP-A51A08/2020\|EPI_ISL_598170\|20 | ···················· | ···················· | ····················· | ···················· | ···················· | ···················· |

| | | | | | | | |
|---|---|---|---|---|---|---|---|
| N | 3108 hCoV-19/England/ALDP-A51ACC/2020\|EPI_ISL_598171\|20 | ···················· | ···················· | ···················· | ···················· | ···················· | ···················· |
| N | 3109 hCoV-19/England/ALDP-A51D96/2020\|EPI_ISL_598173\|20 | ···················· | ···················· | ···················· | ···················· | ···················· | ···················· |
| N | 3110 hCoV-19/England/ALDP-A523A2/2020\|EPI_ISL_598174\|20 | ···················· | ···················· | ···················· | ···················· | ···················· | ···················· |
| N | 3111 hCoV-19/England/ALDP-A5169B/2020\|EPI_ISL_598175\|20 | ···················· | ···················· | ···················· | ···················· | ···················· | ···················· |
| N | 3112 hCoV-19/England/ALDP-A52533/2020\|EPI_ISL_598176\|20 | ···················· | ···················· | ···················· | ···················· | ···················· | ···················· |
| N | 3113 hCoV-19/England/ALDP-A5117C/2020\|EPI_ISL_598177\|20 | ···················· | ···················· | ···················· | ···················· | ···················· | ···················· |
| N | 3114 hCoV-19/England/ALDP-A51C8A/2020\|EPI_ISL_598179\|20 | ···················· | ···················· | ···················· | ···················· | ···················· | ···················· |
| N | 3115 hCoV-19/England/ALDP-A51279/2020\|EPI_ISL_598180\|20 | ···················· | ···················· | ···················· | ···················· | ···················· | ···················· |
| N | 3116 hCoV-19/England/ALDP-A528A3/2020\|EPI_ISL_598181\|20 | ···················· | ···················· | ···················· | ···················· | ···················· | ···················· |
| N | 3117 hCoV-19/England/ALDP-A516E6/2020\|EPI_ISL_598182\|20 | ···················· | ···················· | ···················· | ···················· | ···················· | ···················· |
| N | 3118 hCoV-19/England/ALDP-A51B14/2020\|EPI_ISL_598184\|20 | ···················· | ···················· | ···················· | ···················· | ···················· | ···················· |
| N | 3119 hCoV-19/England/ALDP-A511E5/2020\|EPI_ISL_598185\|20 | ···················· | ···················· | ···················· | ···················· | ···················· | ···················· |
| N | 3120 hCoV-19/England/ALDP-A51419/2020\|EPI_ISL_598186\|20 | ···················· | ···················· | ···················· | ···················· | ···················· | ···················· |
| N | 3121 hCoV-19/England/ALDP-A524AF/2020\|EPI_ISL_598187\|20 | ···················· | ···················· | ···················· | ···················· | ···················· | ···················· |
| N | 3122 hCoV-19/England/ALDP-A51A71/2020\|EPI_ISL_598188\|20 | ···················· | ···················· | ···················· | ···················· | ···················· | ···················· |
| N | 3123 hCoV-19/England/ALDP-A51C7B/2020\|EPI_ISL_598189\|20 | ···················· | ···················· | ···················· | ···················· | ···················· | ···················· |
| N | 3124 hCoV-19/England/ALDP-A519DE/2020\|EPI_ISL_598190\|20 | ···················· | ···················· | ···················· | ···················· | ···················· | ···················· |
| N | 3125 hCoV-19/England/ALDP-A523DF/2020\|EPI_ISL_598192\|20 | ···················· | ···················· | ···················· | ···················· | ···················· | ···················· |
| N | 3126 hCoV-19/England/ALDP-A5232A/2020\|EPI_ISL_598193\|20 | ···················· | ···················· | ···················· | ···················· | ···················· | ···················· |
| N | 3127 hCoV-19/England/ALDP-A51DE1/2020\|EPI_ISL_598194\|20 | ···················· | ···················· | ···················· | ···················· | ···················· | ···················· |
| N | 3128 hCoV-19/England/ALDP-A519B0/2020\|EPI_ISL_598195\|20 | ···················· | ·A·················· | ···················· | ···················· | ···················· | ···················· |
| N | 3129 hCoV-19/England/ALDP-A52885/2020\|EPI_ISL_598196\|20 | ···················· | ···················· | ···················· | ···················· | ···················· | ···················· |
| N | 3130 hCoV-19/England/ALDP-A51C20/2020\|EPI_ISL_598197\|20 | ···················· | ···················· | ···················· | ···················· | ···················· | ···················· |
| N | 3131 hCoV-19/England/ALDP-A51516/2020\|EPI_ISL_598198\|20 | ···················· | ···················· | ···················· | ···················· | ···················· | ···················· |
| N | 3132 hCoV-19/England/ALDP-A519CF/2020\|EPI_ISL_598199\|20 | ···················· | ···················· | ···················· | ···················· | ···················· | ···················· |
| N | 3133 hCoV-19/England/ALDP-A512A6/2020\|EPI_ISL_598200\|20 | ···················· | ···················· | ···················· | ···················· | ···················· | ···················· |
| N | 3134 hCoV-19/England/ALDP-A51FEB/2020\|EPI_ISL_598201\|20 | ···················· | ···················· | ···················· | ···················· | ···················· | ···················· |
| N | 3135 hCoV-19/England/ALDP-A51C4E/2020\|EPI_ISL_598202\|20 | ···················· | ···················· | ···················· | ···················· | ···················· | ···················· |
| N | 3136 hCoV-19/England/ALDP-A513B2/2020\|EPI_ISL_598203\|20 | ···················· | ···················· | ···················· | ···················· | ···················· | ···················· |
| N | 3137 hCoV-19/England/ALDP-A527C4/2020\|EPI_ISL_598230\|20 | ···················· | ···················· | ···················· | ···················· | ···················· | ···················· |
| N | 3138 hCoV-19/England/ALDP-A51A17/2020\|EPI_ISL_598231\|20 | ···················· | ···················· | ···················· | ···················· | ···················· | ···················· |
| N | 3139 hCoV-19/England/ALDP-A52445/2020\|EPI_ISL_598232\|20 | ···················· | ···················· | ···················· | ···················· | ···················· | ···················· |
| N | 3140 hCoV-19/England/QEUH-A0BB64/2020\|EPI_ISL_599458\|20 | ···················· | ···················· | ···················· | ···················· | ···················· | ···················· |
| N | 3141 hCoV-19/England/QEUH-A0BA3A/2020\|EPI_ISL_599459\|20 | ···················· | ···················· | ···················· | ···················· | ···················· | ···················· |
| N | 3142 hCoV-19/England/QEUH-A0B34E/2020\|EPI_ISL_599460\|20 | ···················· | ···················· | ···················· | ···················· | ···················· | ···················· |
| N | 3143 hCoV-19/England/QEUH-A0BC25/2020\|EPI_ISL_599461\|20 | ···················· | ···················· | ···················· | ···················· | ···················· | ···················· |
| N | 3144 hCoV-19/England/QEUH-A0B9A6/2020\|EPI_ISL_599462\|20 | ···················· | ···················· | ···················· | ···················· | ···················· | ···················· |
| N | 3145 hCoV-19/England/QEUH-A0BE10/2020\|EPI_ISL_599464\|20 | ···················· | ···················· | ···················· | ···················· | ···················· | ···················· |
| N | 3146 hCoV-19/England/QEUH-A0B478/2020\|EPI_ISL_599465\|20 | ···················· | ···················· | ···················· | ···················· | ···················· | ···················· |
| N | 3147 hCoV-19/Scotland/QEUH-A0B7F7/2020\|EPI_ISL_599466\|2 | ···················· | ···················· | ···················· | ···················· | ···················· | ···················· |
| N | 3148 hCoV-19/England/QEUH-A0B751/2020\|EPI_ISL_599467\|20 | ···················· | ······C············· | ···················· | ···················· | ···················· | ···················· |
| N | 3149 hCoV-19/Scotland/QEUH-A0B85E/2020\|EPI_ISL_599468\|2 | ···················· | ···················· | ···················· | ···················· | ···················· | ···················· |
| N | 3150 hCoV-19/England/QEUH-A0B37B/2020\|EPI_ISL_599469\|20 | ···················· | ···················· | ···················· | ···················· | ···················· | ···················· |
| N | 3151 hCoV-19/England/QEUH-A0BA2B/2020\|EPI_ISL_599470\|20 | ···················· | ···················· | ···················· | ···················· | ···················· | ···················· |
| N | 3152 hCoV-19/England/QEUH-A0BA1C/2020\|EPI_ISL_599471\|20 | ···················· | ···················· | ···················· | ···················· | ···················· | ···················· |
| N | 3153 hCoV-19/Scotland/QEUH-A0BD13/2020\|EPI_ISL_599472\|2 | ···················· | ···················· | ···················· | ···················· | ···················· | ···················· |
| N | 3154 hCoV-19/England/QEUH-A0BDD7/2020\|EPI_ISL_599473\|20 | ···················· | ···················· | ···················· | ···················· | ···················· | ···················· |
| N | 3155 hCoV-19/England/QEUH-A0B43C/2020\|EPI_ISL_599475\|20 | ···················· | ···················· | ···················· | ···················· | ···················· | ···················· |
| N | 3156 hCoV-19/England/QEUH-A0B654/2020\|EPI_ISL_599476\|20 | ···················· | ···················· | ···················· | ···················· | ···················· | ···················· |
| N | 3157 hCoV-19/Scotland/QEUH-A0B548/2020\|EPI_ISL_599477\|2 | ···················· | ···················· | ···················· | ···················· | ···················· | ···················· |
| N | 3158 hCoV-19/England/QEUH-A0B19F/2020\|EPI_ISL_599479\|20 | ···················· | ···················· | ···················· | ···················· | ···················· | ···················· |
| N | 3159 hCoV-19/Scotland/QEUH-A0B487/2020\|EPI_ISL_599480\|2 | ···················· | ···················· | ···················· | ···················· | ···················· | ···················· |
| N | 3160 hCoV-19/England/QEUH-A0B6AF/2020\|EPI_ISL_599481\|20 | ···················· | ···················· | ···················· | ···················· | ···················· | ···················· |
| N | 3161 hCoV-19/England/QEUH-A0AD05/2020\|EPI_ISL_599482\|20 | ···················· | ···················· | ···················· | ···················· | ····T··············· | ···················· |
| N | 3162 hCoV-19/England/QEUH-9DE0B7/2020\|EPI_ISL_599485\|20 | ···················· | ···················· | ···················· | ···················· | ···················· | ···················· |
| N | 3163 hCoV-19/England/QEUH-9DE62E/2020\|EPI_ISL_599487\|20 | ···················· | ···················· | ···················· | ···················· | ···················· | ···················· |
| N | 3164 hCoV-19/England/QEUH-9DDE90/2020\|EPI_ISL_599488\|20 | ···················· | ···················· | ···················· | ···················· | ······T············· | ···················· |
| N | 3165 hCoV-19/Scotland/QEUH-9DD9F9/2020\|EPI_ISL_599489\|2 | ···················· | ···················· | ···················· | ···················· | ···················· | ···················· |
| N | 3166 hCoV-19/Scotland/QEUH-9DD513/2020\|EPI_ISL_599491\|2 | ···················· | ···················· | ···················· | ···················· | ···················· | ···················· |
| N | 3167 hCoV-19/Scotland/QEUH-9DD856/2020\|EPI_ISL_599492\|2 | ···················· | ···················· | ···················· | ···················· | ···················· | ···················· |
| N | 3168 hCoV-19/England/QEUH-9DE66A/2020\|EPI_ISL_599493\|20 | ···················· | ···················· | ···················· | ···················· | ···················· | ···················· |
| N | 3169 hCoV-19/England/QEUH-9DE239/2020\|EPI_ISL_599494\|20 | ···················· | ···················· | ···················· | ···················· | ···················· | ···················· |
| N | 3170 hCoV-19/England/QEUH-9DE48E/2020\|EPI_ISL_599495\|20 | ···················· | ·A·················· | ···················· | ···················· | ···················· | ··A················· |
| N | 3171 hCoV-19/England/QEUH-9DCDB2/2020\|EPI_ISL_599496\|20 | ···················· | ·A·················· | ···················· | ···················· | ···················· | ··A················· |
| N | 3172 hCoV-19/England/QEUH-9DCF52/2020\|EPI_ISL_599497\|20 | ···················· | ·A·················· | ···················· | ···················· | ···················· | ··A················· |
| N | 3173 hCoV-19/England/QEUH-9DE0D5/2020\|EPI_ISL_599498\|20 | ···················· | ···················· | ···················· | ···················· | ···················· | ···················· |
| N | 3174 hCoV-19/England/QEUH-9DD9BD/2020\|EPI_ISL_599499\|20 | ···················· | ···················· | ···················· | ···················· | ···················· | ···················· |
| N | 3175 hCoV-19/England/QEUH-9DDCD2/2020\|EPI_ISL_599500\|20 | ···················· | ···················· | ···················· | ···················· | ···················· | ···················· |
| N | 3176 hCoV-19/England/QEUH-9DDB20/2020\|EPI_ISL_599501\|20 | ···················· | ···················· | ···················· | ···················· | ···················· | ···················· |
| N | 3177 hCoV-19/England/QEUH-9DD443/2020\|EPI_ISL_599503\|20 | ···················· | ···················· | ···················· | ···················· | ···················· | ···················· |
| N | 3178 hCoV-19/England/QEUH-9DCCB5/2020\|EPI_ISL_599504\|20 | ···················· | ···················· | ···················· | ···················· | ···················· | ···················· |
| N | 3179 hCoV-19/England/QEUH-9DE12D/2020\|EPI_ISL_599505\|20 | ···················· | ·A·················· | ···················· | ···················· | ···················· | ···················· |
| N | 3180 hCoV-19/England/QEUH-9DE11E/2020\|EPI_ISL_599506\|20 | ···················· | ·A·················· | ···················· | ···················· | ···················· | ···················· |
| N | 3181 hCoV-19/Scotland/QEUH-9DD66B/2020\|EPI_ISL_599508\|2 | ···················· | ···················· | ···················· | ···················· | ···················· | ···················· |
| N | 3182 hCoV-19/England/QEUH-9DE381/2020\|EPI_ISL_599509\|20 | ···················· | ···················· | ···················· | ···················· | ···················· | ···················· |
| N | 3183 hCoV-19/England/QEUH-9DCCA6/2020\|EPI_ISL_599510\|20 | ···················· | ·A·················· | ···················· | ···················· | ···················· | ··A················· |
| N | 3184 hCoV-19/Scotland/QEUH-9DE04E/2020\|EPI_ISL_599511\|2 | ···················· | ···················· | ···················· | ···················· | ···················· | ···················· |
| N | 3185 hCoV-19/England/QEUH-9DDE72/2020\|EPI_ISL_599512\|20 | ···················· | ···················· | ···················· | ···················· | ···················· | ···················· |
| N | 3186 hCoV-19/England/QEUH-9DD874/2020\|EPI_ISL_599513\|20 | ···················· | ···················· | ···················· | ···················· | ···················· | ···················· |
| N | 3187 hCoV-19/England/QEUH-9DE4E8/2020\|EPI_ISL_599514\|20 | ···················· | ···················· | ···················· | ···················· | ···················· | ···················· |

| | | | | | | | | |
|---|---|---|---|---|---|---|---|---|
| N | 3188 | hCoV-19/England/QEUH-9DD971/2020\|EPI_ISL_599517\|20 | ···················· | ···················· | ···················· | ···················· | ···T················ | ···················· |
| N | 3189 | hCoV-19/England/QEUH-9DCE64/2020\|EPI_ISL_599518\|20 | ···················· | ···················· | ···················· | ···················· | ···················· | ···················· |
| N | 3190 | hCoV-19/England/QEUH-9DDDFD/2020\|EPI_ISL_599519\|20 | ···················· | ···················· | ···················· | ···················· | ···················· | ···················· |
| N | 3191 | hCoV-19/England/QEUH-9DD012/2020\|EPI_ISL_599520\|20 | ···················· | ···················· | ···················· | ···················· | ···················· | ···················· |
| N | 3192 | hCoV-19/Scotland/QEUH-9DD795/2020\|EPI_ISL_599521\|2 | ···················· | ···················· | ···················· | ···················· | ···················· | ···················· |
| N | 3193 | hCoV-19/England/QEUH-9DD9DB/2020\|EPI_ISL_599522\|20 | ···················· | ···················· | ···················· | ···················· | ···················· | ···················· |
| N | 3194 | hCoV-19/England/QEUH-9DD908/2020\|EPI_ISL_599523\|20 | ···················· | ···················· | ···················· | ···················· | ···················· | ···················· |
| N | 3195 | hCoV-19/Scotland/QEUH-9DD285/2020\|EPI_ISL_599525\|2 | ···················· | ···················· | ···················· | ···················· | ·····T·············· | ···················· |
| N | 3196 | hCoV-19/England/QEUH-9DCE46/2020\|EPI_ISL_599526\|20 | ···················· | ···················· | ···················· | ···················· | ···················· | ···················· |
| N | 3197 | hCoV-19/England/QEUH-9DE2FD/2020\|EPI_ISL_599528\|20 | ···················· | ···················· | ···················· | ···················· | ···················· | ···················· |
| N | 3198 | hCoV-19/England/QEUH-9DD71D/2020\|EPI_ISL_599529\|20 | ···················· | ···················· | ···················· | ···················· | ···T················ | ···················· |
| N | 3199 | hCoV-19/England/QEUH-9DD953/2020\|EPI_ISL_599530\|20 | ···················· | ···················· | ···················· | ···················· | ···················· | ···················· |
| N | 3200 | hCoV-19/England/QEUH-9DD425/2020\|EPI_ISL_599531\|20 | ···················· | ···················· | ···················· | ···················· | ···················· | ···················· |
| N | 3201 | hCoV-19/England/QEUH-9DDA7E/2020\|EPI_ISL_599532\|20 | ···················· | ···················· | ···················· | ···················· | ···················· | ···················· |
| N | 3202 | hCoV-19/England/QEUH-9DDFAC/2020\|EPI_ISL_599533\|20 | ···················· | ···················· | ···················· | ···················· | ···················· | ···················· |
| N | 3203 | hCoV-19/England/QEUH-9DE521/2020\|EPI_ISL_599534\|20 | ···················· | ·A·················· | ···················· | ···················· | ···················· | ··A················· |
| N | 3204 | hCoV-19/England/QEUH-9DD09A/2020\|EPI_ISL_599561\|20 | ···················· | ···················· | ···················· | ···················· | ···················· | ···················· |
| N | 3205 | hCoV-19/England/QEUH-9DDC96/2020\|EPI_ISL_599562\|20 | ···················· | ···················· | ···················· | ···················· | ···················· | ···················· |
| N | 3206 | hCoV-19/England/QEUH-9DE8DD/2020\|EPI_ISL_599563\|20 | ···················· | ·A·················· | ···················· | ···················· | ···················· | ···················· |
| N | 3207 | hCoV-19/Scotland/QEUH-9F7D66/2020\|EPI_ISL_600918\|2 | ···················· | ···················· | ···················· | ···················· | ···················· | ···················· |
| N | 3208 | hCoV-19/Scotland/QEUH-9F94F6/2020\|EPI_ISL_600919\|2 | ···················· | ···················· | ···················· | ···················· | ···················· | ···················· |
| N | 3209 | hCoV-19/Scotland/QEUH-9E4203/2020\|EPI_ISL_600954\|2 | ···················· | ···················· | ···················· | ···················· | ···················· | ···················· |
| N | 3210 | hCoV-19/Scotland/QEUH-9E4CF6/2020\|EPI_ISL_600955\|2 | ···················· | ···················· | ···················· | ···················· | ···················· | ···················· |
| N | 3211 | hCoV-19/Scotland/QEUH-9E4E96/2020\|EPI_ISL_600957\|2 | ···················· | ···················· | ···················· | ···················· | ···················· | ···················· |
| N | 3212 | hCoV-19/Scotland/QEUH-9E429A/2020\|EPI_ISL_600958\|2 | ···················· | ···················· | ···················· | ···················· | ···················· | ···················· |
| N | 3213 | hCoV-19/Scotland/QEUH-9E4F1B/2020\|EPI_ISL_600959\|2 | ···················· | ···················· | ···················· | ···················· | ···················· | ···················· |
| N | 3214 | hCoV-19/Scotland/QEUH-9E4573/2020\|EPI_ISL_600960\|2 | ···················· | ···················· | ···················· | ···················· | ···················· | ···················· |
| N | 3215 | hCoV-19/Scotland/QEUH-9E428B/2020\|EPI_ISL_600961\|2 | ···················· | ···················· | ···················· | ···················· | ···················· | ···················· |
| N | 3216 | hCoV-19/Scotland/QEUH-9E4713/2020\|EPI_ISL_600962\|2 | ···················· | ···················· | ···················· | ···················· | ···················· | ···················· |
| N | 3217 | hCoV-19/Scotland/QEUH-9E42A9/2020\|EPI_ISL_600963\|2 | ···················· | ···················· | ···················· | ···················· | ···················· | ···················· |
| N | 3218 | hCoV-19/Scotland/QEUH-9E46DA/2020\|EPI_ISL_600964\|2 | ···················· | ···················· | ···················· | ···················· | ···················· | ···················· |
| N | 3219 | hCoV-19/Scotland/QEUH-9E4528/2020\|EPI_ISL_600965\|2 | ···················· | ···················· | ···················· | ···················· | ···················· | ···················· |
| N | 3220 | hCoV-19/Scotland/QEUH-9E4494/2020\|EPI_ISL_600966\|2 | ···················· | ···················· | ···················· | ···················· | ···················· | ···················· |
| N | 3221 | hCoV-19/Scotland/QEUH-9E44D0/2020\|EPI_ISL_600969\|2 | ···················· | ···················· | ···················· | ···················· | ···················· | ···················· |
| N | 3222 | hCoV-19/Scotland/QEUH-9E47F5/2020\|EPI_ISL_600970\|2 | ···················· | ···················· | ···················· | ···················· | ···················· | ···················· |
| N | 3223 | hCoV-19/Scotland/QEUH-9E484D/2020\|EPI_ISL_600971\|2 | ···················· | ···················· | ···················· | ···················· | ···················· | ···················· |
| N | 3224 | hCoV-19/Scotland/QEUH-9E46CB/2020\|EPI_ISL_600972\|2 | ···················· | ···················· | ···················· | ···················· | ···················· | ···················· |
| N | 3225 | hCoV-19/Scotland/QEUH-9E5387/2020\|EPI_ISL_600973\|2 | ···················· | ···················· | ···················· | ···················· | ···················· | ···················· |
| N | 3226 | hCoV-19/Scotland/QEUH-9E4B17/2020\|EPI_ISL_600975\|2 | ···················· | ···················· | ···················· | ···················· | ···················· | ···················· |
| N | 3227 | hCoV-19/Scotland/QEUH-9E4625/2020\|EPI_ISL_600976\|2 | ···················· | ···················· | ···················· | ···················· | ···················· | ···················· |
| N | 3228 | hCoV-19/Scotland/QEUH-9E44C1/2020\|EPI_ISL_600977\|2 | ···················· | ···················· | ···················· | ···················· | ···················· | ···················· |
| N | 3229 | hCoV-19/Scotland/QEUH-9E51BA/2020\|EPI_ISL_600978\|2 | ···················· | ···················· | ···················· | ···················· | ···················· | ···················· |
| N | 3230 | hCoV-19/Scotland/QEUH-9E4A29/2020\|EPI_ISL_600979\|2 | ···················· | ···················· | ···················· | ···················· | ···················· | ···················· |
| N | 3231 | hCoV-19/Scotland/QEUH-9E4810/2020\|EPI_ISL_600980\|2 | ···················· | ···················· | ···················· | ···················· | ···················· | ···················· |
| N | 3232 | hCoV-19/Scotland/QEUH-9E487A/2020\|EPI_ISL_600982\|2 | ···················· | ···················· | ···················· | ···················· | ···················· | ···················· |
| N | 3233 | hCoV-19/Scotland/QEUH-9E5509/2020\|EPI_ISL_600983\|2 | ···················· | ···················· | ···················· | ···················· | ···················· | ···················· |
| N | 3234 | hCoV-19/Scotland/QEUH-9E4661/2020\|EPI_ISL_600984\|2 | ···················· | ···················· | ···················· | ···················· | ···················· | ···················· |
| N | 3235 | hCoV-19/Scotland/QEUH-9E4555/2020\|EPI_ISL_600985\|2 | ···················· | ···················· | ···················· | ···················· | ···················· | ···················· |
| N | 3236 | hCoV-19/Scotland/QEUH-9E55FA/2020\|EPI_ISL_600987\|2 | ···················· | ···················· | ···················· | ···················· | ···················· | ···················· |
| N | 3237 | hCoV-19/Scotland/QEUH-9E5378/2020\|EPI_ISL_600988\|2 | ···················· | ···················· | ···················· | ···················· | ···················· | ···················· |
| N | 3238 | hCoV-19/Scotland/QEUH-9E4C7E/2020\|EPI_ISL_600989\|2 | ···················· | ···················· | ···················· | ···················· | ···················· | ···················· |
| N | 3239 | hCoV-19/Scotland/QEUH-9E42C7/2020\|EPI_ISL_600990\|2 | ···················· | ···················· | ···················· | ···················· | ···················· | ···················· |
| N | 3240 | hCoV-19/Scotland/QEUH-9E46E9/2020\|EPI_ISL_600991\|2 | ···················· | ···················· | ···················· | ···················· | ···················· | ···················· |
| N | 3241 | hCoV-19/Scotland/QEUH-9E4485/2020\|EPI_ISL_600992\|2 | ···················· | ···················· | ···················· | ···················· | ···················· | ···················· |
| N | 3242 | hCoV-19/Scotland/QEUH-9E52A8/2020\|EPI_ISL_600993\|2 | ···················· | ···················· | ···················· | ···················· | ···················· | ···················· |
| N | 3243 | hCoV-19/Scotland/QEUH-9E4072/2020\|EPI_ISL_600994\|2 | ···················· | ···················· | ···················· | ···················· | ···················· | ···················· |
| N | 3244 | hCoV-19/Scotland/QEUH-9E432E/2020\|EPI_ISL_600995\|2 | ···················· | ···················· | ···················· | ···················· | ···················· | ···················· |
| N | 3245 | hCoV-19/Scotland/QEUH-9E4A74/2020\|EPI_ISL_600996\|2 | ···················· | ···················· | ···················· | ···················· | ···················· | ···················· |
| N | 3246 | hCoV-19/Scotland/QEUH-9E4591/2020\|EPI_ISL_600997\|2 | ···················· | ···················· | ···················· | ···················· | ···················· | ···················· |
| N | 3247 | hCoV-19/England/204191512/2020\|EPI_ISL_603190\|2020 | ···················· | ···················· | ···················· | ···················· | ···················· | ···················· |
| N | 3248 | hCoV-19/England/204191900/2020\|EPI_ISL_603191\|2020 | ···················· | ···················· | ···················· | ···················· | ···················· | ···················· |
| N | 3249 | hCoV-19/Bangladesh/BCSIR-NILMRC_393/2020\|EPI_ISL_6 | ···················· | ···················· | ···················· | ···················· | ···················· | ···················· |
| N | 3250 | hCoV-19/Bangladesh/BCSIR-NILMRC_396/2020\|EPI_ISL_6 | ···················· | ···················· | ···················· | ···················· | ···················· | ···················· |
| N | 3251 | hCoV-19/Bangladesh/BCSIR-NILMRC_397/2020\|EPI_ISL_6 | ···················· | ···················· | ···················· | ···················· | ···················· | ···················· |
| N | 3252 | hCoV-19/Bangladesh/BCSIR-NILMRC_411/2020\|EPI_ISL_6 | ···················· | ···················· | ···················· | ···················· | ···················· | ···················· |
| N | 3253 | hCoV-19/Bangladesh/BCSIR-NILMRC_420/2020\|EPI_ISL_6 | ···················· | ···················· | ···················· | ···················· | ···················· | ···················· |
| N | 3254 | hCoV-19/Bangladesh/BCSIR-NILMRC_421/2020\|EPI_ISL_6 | ···················· | ···················· | ···················· | ···················· | ···················· | ···················· |
| N | 3255 | hCoV-19/Bangladesh/BCSIR-NILMRC_422/2020\|EPI_ISL_6 | ···················· | ···················· | ···················· | ···················· | ···················· | ···················· |
| N | 3256 | hCoV-19/Bangladesh/BCSIR-NILMRC_423/2020\|EPI_ISL_6 | ···················· | ···················· | ···················· | ···················· | ···················· | ···················· |
| N | 3257 | hCoV-19/Bangladesh/BCSIR-NILMRC_424/2020\|EPI_ISL_6 | ···················· | ···················· | ···················· | ···················· | ···················· | ···················· |
| N | 3258 | hCoV-19/Bangladesh/BCSIR-NILMRC_426/2020\|EPI_ISL_6 | ···················· | ···················· | ···················· | ···················· | ···················· | ···················· |
| N | 3259 | hCoV-19/USA/DC-BBI1/2020\|EPI_ISL_603248\|2020-10-02 | ···················· | ···················· | ···················· | ···················· | ···················· | ···················· |
| N | 3260 | hCoV-19/Bangladesh/BCSIR-NILMRC_427/2020\|EPI_ISL_6 | ···················· | ···················· | ···················· | ···················· | ···················· | ···················· |
| N | 3261 | hCoV-19/USA/WA-UW-23621/2020\|EPI_ISL_603251\|2020-0 | ···················· | ···················· | ···················· | ···················· | ···················· | ···················· |
| N | 3262 | hCoV-19/USA/WA-UW-29759/2020\|EPI_ISL_603252\|2020-0 | ···················· | ·A·················· | ···················· | ···················· | ···················· | ···················· |
| N | 3263 | hCoV-19/USA/WA-UW-29740/2020\|EPI_ISL_603254\|2020-0 | ···················· | ·A·················· | ···················· | ···················· | ···················· | ···················· |
| N | 3264 | hCoV-19/USA/WA-UW-29702/2020\|EPI_ISL_603255\|2020-0 | ···················· | ···················· | ···················· | ···················· | ···················· | ···················· |
| N | 3265 | hCoV-19/Switzerland/BE-ETHZ-300234/2020\|EPI_ISL_60 | ···················· | ···················· | ···················· | ···················· | ···················· | ···················· |
| N | 3266 | hCoV-19/Switzerland/ZH-ETHZ-300235/2020\|EPI_ISL_60 | ···················· | ···················· | ···················· | ···················· | ···················· | ···················· |
| N | 3267 | hCoV-19/Switzerland/ZH-ETHZ-300236/2020\|EPI_ISL_60 | ···················· | ···················· | ···················· | ···················· | ···················· | ···················· |

| | | | | | | | | |
|---|---|---|---|---|---|---|---|---|
| N | 3268 | hCoV-19/Switzerland/BL-ETHZ-300237/2020\|EPI_ISL_60 | ···················· | ···················· | ···················· | ···················· | ···················· | ···················· |
| N | 3269 | hCoV-19/Switzerland/GR-ETHZ-300239/2020\|EPI_ISL_60 | ···················· | ···················· | ···················· | ···················· | ···················· | ···················· |
| N | 3270 | hCoV-19/Switzerland/ZH-ETHZ-300240/2020\|EPI_ISL_60 | ···················· | ···················· | ···················· | ···················· | ···················· | ···················· |
| N | 3271 | hCoV-19/Switzerland/GR-ETHZ-300241/2020\|EPI_ISL_60 | ···················· | ···················· | ···················· | ···················· | ···················· | ···················· |
| N | 3272 | hCoV-19/Switzerland/ZH-ETHZ-300242/2020\|EPI_ISL_60 | ···················· | ···················· | ···················· | ···················· | ···················· | ···················· |
| N | 3273 | hCoV-19/Switzerland/SG-ETHZ-300243/2020\|EPI_ISL_60 | ···················· | ···················· | ···················· | ···················· | ···················· | ···················· |
| N | 3274 | hCoV-19/Switzerland/SZ-ETHZ-300244/2020\|EPI_ISL_60 | ···················· | ···················· | ···················· | ···················· | ···················· | ···················· |
| N | 3275 | hCoV-19/Switzerland/SG-ETHZ-300245/2020\|EPI_ISL_60 | ···················· | ···················· | ···················· | ···················· | ···················· | ···················· |
| N | 3276 | hCoV-19/Switzerland/ZH-ETHZ-300246/2020\|EPI_ISL_60 | ···················· | ···················· | ···················· | ···················· | ···················· | ···················· |
| N | 3277 | hCoV-19/Switzerland/VD-ETHZ-300248/2020\|EPI_ISL_60 | ···················· | ···················· | ···················· | ···················· | ···················· | ···················· |
| N | 3278 | hCoV-19/Switzerland/GR-ETHZ-300249/2020\|EPI_ISL_60 | ···················· | ···················· | ···················· | ···················· | ···················· | ···················· |
| N | 3279 | hCoV-19/Switzerland/VD-ETHZ-300250/2020\|EPI_ISL_60 | ···················· | ···················· | ···················· | ···················· | ···················· | ···················· |
| N | 3280 | hCoV-19/Switzerland/VD-ETHZ-300251/2020\|EPI_ISL_60 | ···················· | ···················· | ···················· | ···················· | ···················· | ···················· |
| N | 3281 | hCoV-19/Switzerland/VD-ETHZ-300252/2020\|EPI_ISL_60 | ···················· | ···················· | ···················· | ···················· | ···················· | ···················· |
| N | 3282 | hCoV-19/Switzerland/VD-ETHZ-300256/2020\|EPI_ISL_60 | ···················· | ···················· | ···················· | ···················· | ···················· | ···················· |
| N | 3283 | hCoV-19/Switzerland/AG-ETHZ-300257/2020\|EPI_ISL_60 | ···················· | ···················· | ···················· | ···················· | ···················· | ···················· |
| N | 3284 | hCoV-19/Switzerland/SO-ETHZ-300258/2020\|EPI_ISL_60 | ···················· | ···················· | ···················· | ···················· | ···················· | ···················· |
| N | 3285 | hCoV-19/Switzerland/ZH-ETHZ-300259/2020\|EPI_ISL_60 | ···················· | ···················· | ···················· | ···················· | ···················· | ···················· |
| N | 3286 | hCoV-19/Switzerland/ZH-ETHZ-300260/2020\|EPI_ISL_60 | ···················· | ···················· | ···················· | ···················· | ···················· | ···················· |
| N | 3287 | hCoV-19/Switzerland/ZH-ETHZ-300261/2020\|EPI_ISL_60 | ···················· | ···················· | ···················· | ···················· | ···················· | ···················· |
| N | 3288 | hCoV-19/Switzerland/ZH-ETHZ-300262/2020\|EPI_ISL_60 | ···················· | ···················· | ···················· | ···················· | ···················· | ···················· |
| N | 3289 | hCoV-19/Switzerland/VD-ETHZ-300263/2020\|EPI_ISL_60 | ···················· | ···················· | ···················· | ···················· | ···················· | ···················· |
| N | 3290 | hCoV-19/Switzerland/BE-ETHZ-300264/2020\|EPI_ISL_60 | ···················· | ···················· | ···················· | ···················· | ···················· | ···················· |
| N | 3291 | hCoV-19/Switzerland/UR-ETHZ-300265/2020\|EPI_ISL_60 | ···················· | ···················· | ···················· | ···················· | ···················· | ···················· |
| N | 3292 | hCoV-19/Switzerland/VD-ETHZ-300266/2020\|EPI_ISL_60 | ···················· | ···················· | ···················· | ···················· | ···················· | ···················· |
| N | 3293 | hCoV-19/Switzerland/VD-ETHZ-300267/2020\|EPI_ISL_60 | ···················· | ···················· | ···················· | ···················· | ···················· | ···················· |
| N | 3294 | hCoV-19/USA/NJ-QDX-2273/2020\|EPI_ISL_604560\|2020-0 | ···················· | ···················· | ···················· | ···················· | ···················· | ···················· |
| N | 3295 | hCoV-19/USA/NJ-QDX-2278/2020\|EPI_ISL_604561\|2020-0 | ···················· | ···················· | ···················· | ···················· | ···················· | ···················· |
| N | 3296 | hCoV-19/USA/NJ-QDX-2288/2020\|EPI_ISL_604563\|2020-0 | ···················· | ···················· | ···················· | ···················· | ···················· | ···················· |
| N | 3297 | hCoV-19/USA/NJ-QDX-2294/2020\|EPI_ISL_604564\|2020-0 | ···················· | ···················· | ···················· | ···················· | ···················· | ···················· |
| N | 3298 | hCoV-19/USA/NY-QDX-2188/2020\|EPI_ISL_604565\|2020-0 | ···················· | ···················· | ···················· | ···················· | ···················· | ···················· |
| N | 3299 | hCoV-19/USA/NY-QDX-2198/2020\|EPI_ISL_604566\|2020-0 | ···················· | ···················· | ···················· | ···················· | ···················· | ···················· |
| N | 3300 | hCoV-19/USA/NY-QDX-2222/2020\|EPI_ISL_604567\|2020-0 | ···················· | ···················· | ···················· | ···················· | ···················· | ···················· |
| N | 3301 | hCoV-19/USA/NY-QDX-2227/2020\|EPI_ISL_604568\|2020-0 | ···················· | ···················· | ···················· | ···················· | ···················· | ···················· |
| N | 3302 | hCoV-19/USA/NY-QDX-2228/2020\|EPI_ISL_604569\|2020-0 | ···················· | ···················· | ···················· | ···················· | ···················· | ···················· |
| N | 3303 | hCoV-19/USA/NY-QDX-2231/2020\|EPI_ISL_604570\|2020-0 | ···················· | ···················· | ···················· | ···················· | ···················· | ···················· |
| N | 3304 | hCoV-19/USA/NY-QDX-2237/2020\|EPI_ISL_604571\|2020-0 | ···················· | ···················· | ···················· | ···················· | ···················· | ···················· |
| N | 3305 | hCoV-19/USA/NY-QDX-2243/2020\|EPI_ISL_604572\|2020-0 | ···················· | ···················· | ···················· | ···················· | ···················· | ···················· |
| N | 3306 | hCoV-19/USA/NY-QDX-2266/2020\|EPI_ISL_604573\|2020-0 | ···················· | ···················· | ···················· | ···················· | ······A············· | ···················· |
| N | 3307 | hCoV-19/USA/NY-QDX-2292/2020\|EPI_ISL_604575\|2020-0 | ···················· | ···················· | ···················· | ···················· | ···················· | ···················· |
| N | 3308 | hCoV-19/USA/NY-QDX-2293/2020\|EPI_ISL_604576\|2020-0 | ···················· | ···················· | ···················· | ···················· | ···················· | ···················· |
| N | 3309 | hCoV-19/USA/NY-QDX-2304/2020\|EPI_ISL_604577\|2020-0 | ···················· | ···················· | ···················· | ···················· | ···················· | ···················· |
| N | 3310 | hCoV-19/USA/UT-QDX-2189/2020\|EPI_ISL_604578\|2020-0 | ···················· | ···················· | ···················· | ···················· | ···················· | ···················· |
| N | 3311 | hCoV-19/USA/UT-QDX-2192/2020\|EPI_ISL_604579\|2020-0 | ···················· | ···················· | ···················· | ···················· | ···················· | ···················· |
| N | 3312 | hCoV-19/USA/UT-QDX-2194/2020\|EPI_ISL_604580\|2020-0 | ···················· | ···················· | ···················· | ···················· | ···················· | ···················· |
| N | 3313 | hCoV-19/USA/UT-QDX-2201/2020\|EPI_ISL_604581\|2020-0 | ···················· | ···················· | ···················· | ···················· | ···················· | ···················· |
| N | 3314 | hCoV-19/USA/UT-QDX-2202/2020\|EPI_ISL_604582\|2020-0 | ···················· | ···················· | ···················· | ···················· | ···················· | ···················· |
| N | 3315 | hCoV-19/USA/UT-QDX-2207/2020\|EPI_ISL_604583\|2020-0 | ···················· | ···················· | ···················· | ···················· | ···················· | ···················· |
| N | 3316 | hCoV-19/USA/UT-QDX-2208/2020\|EPI_ISL_604584\|2020-0 | ···················· | ···················· | ···················· | ···················· | ···················· | ···················· |
| N | 3317 | hCoV-19/USA/UT-QDX-2213/2020\|EPI_ISL_604585\|2020-0 | ···················· | ···················· | ···················· | ···················· | ···················· | ···················· |
| N | 3318 | hCoV-19/USA/UT-QDX-2214/2020\|EPI_ISL_604586\|2020-0 | ···················· | ···················· | ···················· | ···················· | ···················· | ···················· |
| N | 3319 | hCoV-19/USA/UT-QDX-2217/2020\|EPI_ISL_604588\|2020-0 | ···················· | ···················· | ···················· | ···················· | ···················· | ···················· |
| N | 3320 | hCoV-19/USA/UT-QDX-2224/2020\|EPI_ISL_604590\|2020-0 | ···················· | ···················· | ···················· | ···················· | ···················· | ···················· |
| N | 3321 | hCoV-19/USA/UT-QDX-2230/2020\|EPI_ISL_604591\|2020-0 | ···················· | ···················· | ···················· | ···················· | ···················· | ···················· |
| N | 3322 | hCoV-19/USA/UT-QDX-2233/2020\|EPI_ISL_604592\|2020-0 | ···················· | ···················· | ···················· | ···················· | ···················· | ···················· |
| N | 3323 | hCoV-19/USA/UT-QDX-2234/2020\|EPI_ISL_604593\|2020-0 | ···················· | ···················· | ···················· | ···················· | ···················· | ···················· |
| N | 3324 | hCoV-19/USA/UT-QDX-2238/2020\|EPI_ISL_604594\|2020-0 | ···················· | ···················· | ···················· | ···················· | ···················· | ···················· |
| N | 3325 | hCoV-19/USA/UT-QDX-2240/2020\|EPI_ISL_604595\|2020-0 | ···················· | ···················· | ···················· | ···················· | ···················· | ···················· |
| N | 3326 | hCoV-19/USA/UT-QDX-2247/2020\|EPI_ISL_604597\|2020-0 | ···················· | ···················· | ···················· | ···················· | ···················· | ···················· |
| N | 3327 | hCoV-19/USA/UT-QDX-2248/2020\|EPI_ISL_604598\|2020-0 | ···················· | ···················· | ···················· | ···················· | ···················· | ···················· |
| N | 3328 | hCoV-19/USA/UT-QDX-2249/2020\|EPI_ISL_604599\|2020-0 | ···················· | ···················· | ···················· | ···················· | ···················· | ···················· |
| N | 3329 | hCoV-19/England/ALDP-A52490/2020\|EPI_ISL_598031\|20 | ···················· | ···················· | ···················· | ···················· | ···················· | ···················· |
| N | 3330 | hCoV-19/England/ALDP-A51710/2020\|EPI_ISL_598032\|20 | ···················· | ···················· | ···················· | ···················· | ···················· | ···················· |
| N | 3331 | hCoV-19/England/ALDP-A51F72/2020\|EPI_ISL_598033\|20 | ···················· | ···················· | ···················· | ···················· | ···················· | ···················· |
| N | 3332 | hCoV-19/England/ALDP-A5290A/2020\|EPI_ISL_598034\|20 | ···················· | ···················· | ···················· | ···················· | ···················· | ···················· |
| N | 3333 | hCoV-19/England/ALDP-A527D3/2020\|EPI_ISL_598035\|20 | ···················· | ···················· | ···················· | ···················· | ···················· | ···················· |
| N | 3334 | hCoV-19/England/ALDP-A5115E/2020\|EPI_ISL_598036\|20 | ···················· | ···················· | ···················· | ···················· | ···················· | ···················· |
| N | 3335 | hCoV-19/England/ALDP-A51ADB/2020\|EPI_ISL_598037\|20 | ···················· | ···················· | ···················· | ···················· | ···················· | ···················· |
| N | 3336 | hCoV-19/England/ALDP-A51491/2020\|EPI_ISL_598038\|20 | ···················· | ·A·················· | ···················· | ···················· | ···················· | ···················· |
| N | 3337 | hCoV-19/England/ALDP-A521E4/2020\|EPI_ISL_598090\|20 | ···················· | ···················· | ···················· | ···················· | ···················· | ···················· |
| N | 3338 | hCoV-19/England/ALDP-A52014/2020\|EPI_ISL_598091\|20 | ···················· | ···················· | ···················· | ···················· | ···················· | ···················· |
| N | 3339 | hCoV-19/England/ALDP-A522F0/2020\|EPI_ISL_598092\|20 | ···················· | ···················· | ···················· | ···················· | ···················· | ···················· |
| N | 3340 | hCoV-19/England/ALDP-A51EC0/2020\|EPI_ISL_598093\|20 | ···················· | ···················· | ···················· | ···················· | ···················· | ···················· |
| N | 3341 | hCoV-19/England/ALDP-A5222D/2020\|EPI_ISL_598095\|20 | ···················· | ···················· | ···················· | ···················· | ···················· | ···················· |
| N | 3342 | hCoV-19/England/ALDP-A51F18/2020\|EPI_ISL_598096\|20 | ···················· | ···················· | ···················· | ···················· | ···················· | ···················· |
| N | 3343 | hCoV-19/England/ALDP-A522B4/2020\|EPI_ISL_598097\|20 | ···················· | ···················· | ···················· | ···················· | ···················· | ···················· |
| N | 3344 | hCoV-19/England/ALDP-A520F6/2020\|EPI_ISL_598098\|20 | ···················· | ···················· | ···················· | ···················· | ······T············· | ···················· |
| N | 3345 | hCoV-19/England/ALDP-A5176B/2020\|EPI_ISL_598099\|20 | ···················· | ···················· | ···················· | ···················· | ···················· | ···················· |
| N | 3346 | hCoV-19/Wales/ALDP-A528D0/2020\|EPI_ISL_598204\|2020 | ···················· | ···················· | ···················· | ···················· | ···················· | ···················· |
| N | 3347 | hCoV-19/England/ALDP-A51AEA/2020\|EPI_ISL_598205\|20 | ···················· | ···················· | ···················· | ···················· | ···················· | ···················· |

| | | | | | | | | |
|---|---|---|---|---|---|---|---|---|
| N | 3348 | hCoV-19/England/ALDP-A523B1/2020\|EPI_ISL_598206\|20 | ···················· | ···················· | ···················· | ···················· | ···················· | ···················· |
| N | 3349 | hCoV-19/England/ALDP-A51F45/2020\|EPI_ISL_598207\|20 | ···················· | ···················· | ···················· | ···················· | ···················· | ···················· |
| N | 3350 | hCoV-19/England/ALDP-A51B7E/2020\|EPI_ISL_598208\|20 | ···················· | ···················· | ···················· | ···················· | ···················· | ···················· |
| N | 3351 | hCoV-19/England/ALDP-A51525/2020\|EPI_ISL_598209\|20 | ···················· | ···················· | ···················· | ···················· | ···················· | ···················· |
| N | 3352 | hCoV-19/England/ALDP-A5109D/2020\|EPI_ISL_598210\|20 | ···················· | ···················· | ···················· | ···················· | ···················· | ···················· |
| N | 3353 | hCoV-19/England/ALDP-A52937/2020\|EPI_ISL_598211\|20 | ···················· | ···················· | ···················· | ···················· | ···················· | ···················· |
| N | 3354 | hCoV-19/England/ALDP-A51EB1/2020\|EPI_ISL_598212\|20 | ···················· | ···················· | ···················· | ···················· | ···················· | ···················· |
| N | 3355 | hCoV-19/England/ALDP-A51F36/2020\|EPI_ISL_598214\|20 | ···················· | ···················· | ···················· | ···················· | ···················· | ···················· |
| N | 3356 | hCoV-19/England/ALDP-A5122E/2020\|EPI_ISL_598215\|20 | ···················· | ···················· | ···················· | ···················· | ···················· | ···················· |
| N | 3357 | hCoV-19/England/ALDP-A51CC6/2020\|EPI_ISL_598216\|20 | ···················· | ···················· | ···················· | ···················· | ···················· | ···················· |
| N | 3358 | hCoV-19/England/ALDP-A51BE7/2020\|EPI_ISL_598217\|20 | ···················· | ···················· | ···················· | ···················· | ···················· | ···················· |
| N | 3359 | hCoV-19/England/ALDP-A51BF6/2020\|EPI_ISL_598218\|20 | ···················· | ···················· | ···················· | ···················· | ···················· | ···················· |
| N | 3360 | hCoV-19/England/ALDP-A5180E/2020\|EPI_ISL_598219\|20 | ···················· | ···················· | ···················· | ···················· | ···················· | ···················· |
| N | 3361 | hCoV-19/England/ALDP-A51B32/2020\|EPI_ISL_598220\|20 | ···················· | ···················· | ···················· | ···················· | ···················· | ···················· |
| N | 3362 | hCoV-19/England/ALDP-A51BAB/2020\|EPI_ISL_598221\|20 | ···················· | ···················· | ···················· | ···················· | ···················· | ···················· |
| N | 3363 | hCoV-19/England/ALDP-A52894/2020\|EPI_ISL_598222\|20 | ···················· | ···················· | ···················· | ···················· | ···················· | ···················· |
| N | 3364 | hCoV-19/England/ALDP-A517A7/2020\|EPI_ISL_598223\|20 | ···················· | ···················· | ···················· | ···················· | ···················· | ···················· |
| N | 3365 | hCoV-19/England/ALDP-A51886/2020\|EPI_ISL_598224\|20 | ···················· | ···················· | ···················· | ···················· | ···················· | ···················· |
| N | 3366 | hCoV-19/England/ALDP-A5257F/2020\|EPI_ISL_598225\|20 | ···················· | ···················· | ···················· | ···················· | ···················· | ···················· |
| N | 3367 | hCoV-19/England/ALDP-A51992/2020\|EPI_ISL_598226\|20 | ···················· | ···················· | ···················· | ···················· | ···················· | ···················· |
| N | 3368 | hCoV-19/England/ALDP-A51C11/2020\|EPI_ISL_598227\|20 | ···················· | ···················· | ···················· | ···················· | ···················· | ···················· |
| N | 3369 | hCoV-19/England/ALDP-A5132B/2020\|EPI_ISL_598228\|20 | ···················· | ·A·················· | ···················· | ···················· | ···················· | ···················· |
| N | 3370 | hCoV-19/England/ALDP-A51E48/2020\|EPI_ISL_598229\|20 | ···················· | ···················· | ···················· | ···················· | ···················· | ···················· |
| N | 3371 | hCoV-19/England/ALDP-A51868/2020\|EPI_ISL_598233\|20 | ···················· | ···················· | ···················· | ···················· | ···················· | ···················· |
| N | 3372 | hCoV-19/England/ALDP-A517F2/2020\|EPI_ISL_598234\|20 | ···················· | ···················· | ···················· | ···················· | ···················· | ···················· |
| N | 3373 | hCoV-19/England/ALDP-A52551/2020\|EPI_ISL_598235\|20 | ···················· | ···················· | ···················· | ···················· | ···················· | ···················· |
| N | 3374 | hCoV-19/England/ALDP-A526E5/2020\|EPI_ISL_598237\|20 | ···················· | ···················· | ···················· | ···················· | ···················· | ···················· |
| N | 3375 | hCoV-19/England/ALDP-A5230C/2020\|EPI_ISL_598238\|20 | ···················· | ···················· | ···················· | ···················· | ···················· | ···················· |
| N | 3376 | hCoV-19/England/ALDP-A5182C/2020\|EPI_ISL_598239\|20 | ···················· | ···················· | ···················· | ···················· | ···················· | ···················· |
| N | 3377 | hCoV-19/England/ALDP-A52120/2020\|EPI_ISL_598240\|20 | ···················· | ···················· | ···················· | ···················· | ···················· | ···················· |
| N | 3378 | hCoV-19/England/ALDP-A521D5/2020\|EPI_ISL_598241\|20 | ···················· | ·A·················· | ···················· | ···················· | ···················· | ···A················ |
| N | 3379 | hCoV-19/England/ALDP-A511C7/2020\|EPI_ISL_598243\|20 | ···················· | ·A·················· | ···················· | ···················· | ···················· | ···A················ |
| N | 3380 | hCoV-19/England/ALDP-A512B5/2020\|EPI_ISL_598244\|20 | ···················· | ···················· | ···················· | ···················· | ···················· | ···················· |
| N | 3381 | hCoV-19/England/ALDP-A52357/2020\|EPI_ISL_598245\|20 | ···················· | ···················· | ···················· | ···················· | ···················· | ···················· |
| N | 3382 | hCoV-19/England/ALDP-A5124C/2020\|EPI_ISL_598246\|20 | ···················· | ···················· | ···················· | ···················· | ···················· | ···················· |
| N | 3383 | hCoV-19/England/ALDP-A526A9/2020\|EPI_ISL_598247\|20 | ···················· | ···················· | ···················· | ···················· | ···················· | ···················· |
| N | 3384 | hCoV-19/England/QEUH-A0BB82/2020\|EPI_ISL_599363\|20 | ···················· | ···················· | ···················· | ···················· | ···················· | ···················· |
| N | 3385 | hCoV-19/England/QEUH-A0AAE0/2020\|EPI_ISL_599364\|20 | ···················· | ···················· | ···················· | ···················· | ···················· | ···················· |
| N | 3386 | hCoV-19/Wales/QEUH-A0B26F/2020\|EPI_ISL_599365\|2020 | ···················· | ···················· | ···················· | ···················· | ···················· | ···················· |
| N | 3387 | hCoV-19/Scotland/QEUH-A0BD7D/2020\|EPI_ISL_599366\|2 | ···················· | ···················· | ···················· | ···················· | ···················· | ···················· |
| N | 3388 | hCoV-19/Wales/QEUH-A0B29C/2020\|EPI_ISL_599367\|2020 | ···················· | ···················· | ···················· | ···················· | ···················· | ···················· |
| N | 3389 | hCoV-19/England/QEUH-A0B171/2020\|EPI_ISL_599368\|20 | ···················· | ···················· | ···················· | ···················· | ···················· | ···················· |
| N | 3390 | hCoV-19/Scotland/QEUH-A0B6DC/2020\|EPI_ISL_599369\|2 | ···················· | ···················· | ···················· | ···················· | ···················· | ···················· |
| N | 3391 | hCoV-19/England/QEUH-9DE4D9/2020\|EPI_ISL_599535\|20 | ···················· | ···················· | ···················· | ···················· | ···················· | ···················· |
| N | 3392 | hCoV-19/Scotland/QEUH-9DD80B/2020\|EPI_ISL_599537\|2 | ···················· | ···················· | ···················· | ···················· | ···················· | ···················· |
| N | 3393 | hCoV-19/Scotland/QEUH-9DD470/2020\|EPI_ISL_599539\|2 | ···················· | ···················· | ···················· | ···················· | ···················· | ···················· |
| N | 3394 | hCoV-19/Scotland/QEUH-9DD698/2020\|EPI_ISL_599540\|2 | ···················· | ···················· | ···················· | ···················· | ···················· | ···················· |
| N | 3395 | hCoV-19/England/QEUH-9DE56D/2020\|EPI_ISL_599541\|20 | ···················· | ···················· | ···················· | ···················· | ····T··············· | ···················· |
| N | 3396 | hCoV-19/England/QEUH-9DE002/2020\|EPI_ISL_599542\|20 | ···················· | ···················· | ···················· | ···················· | ···················· | ···················· |
| N | 3397 | hCoV-19/England/QEUH-9DDECD/2020\|EPI_ISL_599543\|20 | ···················· | ···················· | ···················· | ···················· | ····T··············· | ···················· |
| N | 3398 | hCoV-19/England/QEUH-9DD9AE/2020\|EPI_ISL_599544\|20 | ···················· | ···················· | ···················· | ···················· | ···················· | ···················· |
| N | 3399 | hCoV-19/England/QEUH-9DDE54/2020\|EPI_ISL_599545\|20 | ···················· | ···················· | ···················· | ···················· | ···················· | ···················· |
| N | 3400 | hCoV-19/Scotland/QEUH-9DD3BF/2020\|EPI_ISL_599546\|2 | ···················· | ···················· | ···················· | ···················· | ···················· | ···················· |
| N | 3401 | hCoV-19/England/QEUH-9DE336/2020\|EPI_ISL_599547\|20 | ···················· | ·A·················· | ···················· | ···················· | ···················· | ···A················ |
| N | 3402 | hCoV-19/England/QEUH-9DD1A6/2020\|EPI_ISL_599548\|20 | ···················· | ···················· | ···················· | ···················· | ···················· | ···················· |
| N | 3403 | hCoV-19/England/QEUH-9DDA14/2020\|EPI_ISL_599549\|20 | ···················· | ···················· | ···················· | ···················· | ···················· | ···················· |
| N | 3404 | hCoV-19/England/QEUH-9DCFBC/2020\|EPI_ISL_599550\|20 | ···················· | ·A·················· | ···················· | ···················· | ···················· | ···A················ |
| N | 3405 | hCoV-19/England/QEUH-9DD883/2020\|EPI_ISL_599551\|20 | ···················· | ···················· | ···················· | ···················· | ···················· | ···················· |
| N | 3406 | hCoV-19/Scotland/QEUH-9DD3FB/2020\|EPI_ISL_599552\|2 | ···················· | ···················· | ···················· | ···················· | ···················· | ···················· |
| N | 3407 | hCoV-19/England/QEUH-9DCD58/2020\|EPI_ISL_599555\|20 | ···················· | ···················· | ···················· | ···················· | ···················· | ···················· |
| N | 3408 | hCoV-19/Scotland/QEUH-9DD7E0/2020\|EPI_ISL_599557\|2 | ···················· | ···················· | ···················· | ···················· | ···················· | ···················· |
| N | 3409 | hCoV-19/England/QEUH-9DCDA3/2020\|EPI_ISL_599565\|20 | ···················· | ·A·················· | ···················· | ···················· | ···················· | ···················· |
| N | 3410 | hCoV-19/England/QEUH-9DD13D/2020\|EPI_ISL_599566\|20 | ···················· | ·A·················· | ···················· | ···················· | ···················· | ···A················ |
| N | 3411 | hCoV-19/England/QEUH-9DE2DF/2020\|EPI_ISL_599567\|20 | ···················· | ···················· | ···················· | ···················· | ···················· | ···················· |
| N | 3412 | hCoV-19/England/QEUH-9DE2A2/2020\|EPI_ISL_599568\|20 | ···················· | ·A·················· | ···················· | ···················· | ···················· | ···A················ |
| N | 3413 | hCoV-19/England/QEUH-9DDDDF/2020\|EPI_ISL_599569\|20 | ···················· | ·A·················· | ···················· | ···················· | ···················· | ···A················ |
| N | 3414 | hCoV-19/England/QEUH-9DD12E/2020\|EPI_ISL_599570\|20 | ···················· | ···················· | ···················· | ···················· | ···················· | ···················· |
| N | 3415 | hCoV-19/England/QEUH-9DD05E/2020\|EPI_ISL_599571\|20 | ···················· | ·A·················· | ···················· | ···················· | ···················· | ···A················ |
| N | 3416 | hCoV-19/Scotland/QEUH-9DD3A0/2020\|EPI_ISL_599572\|2 | ···················· | ···················· | ···················· | ···················· | ···················· | ···················· |
| N | 3417 | hCoV-19/England/QEUH-9DDA8D/2020\|EPI_ISL_599573\|20 | ···················· | ···················· | ···················· | ···················· | ···················· | ···················· |
| N | 3418 | hCoV-19/England/QEUH-9DE451/2020\|EPI_ISL_599574\|20 | ···················· | ···················· | ···················· | ···················· | ···················· | ···················· |
| N | 3419 | hCoV-19/England/QEUH-9DD6E3/2020\|EPI_ISL_599575\|20 | ···················· | ···················· | ···················· | ···················· | ···················· | ···················· |
| N | 3420 | hCoV-19/Scotland/QEUH-9DD4AD/2020\|EPI_ISL_599578\|2 | ···················· | ···················· | ···················· | ···················· | ···················· | ···················· |
| N | 3421 | hCoV-19/Scotland/QEUH-9F95F3/2020\|EPI_ISL_600910\|2 | ···················· | ···················· | ···················· | ···················· | ···················· | ···················· |
| N | 3422 | hCoV-19/Scotland/QEUH-9F94E7/2020\|EPI_ISL_600911\|2 | ···················· | ···················· | ···················· | ···················· | ···················· | ···················· |
| N | 3423 | hCoV-19/Wales/MILK-9F2378/2020\|EPI_ISL_600912\|2020 | ···················· | ···················· | ···················· | ···················· | ···················· | ···················· |
| N | 3424 | hCoV-19/Wales/MILK-9F2396/2020\|EPI_ISL_600913\|2020 | ···················· | ···················· | ···················· | ···················· | ···················· | ···················· |
| N | 3425 | hCoV-19/Scotland/QEUH-9F7DFD/2020\|EPI_ISL_600914\|2 | ···················· | ···················· | ···················· | ···················· | ···················· | ···················· |
| N | 3426 | hCoV-19/Wales/MILK-9F23C3/2020\|EPI_ISL_600915\|2020 | ···················· | ···················· | ···················· | ···················· | ···················· | ···················· |
| N | 3427 | hCoV-19/Scotland/QEUH-9F74AD/2020\|EPI_ISL_600916\|2 | ···················· | ···················· | ···················· | ···················· | ···················· | ···················· |

| | | | | | | | |
|---|---|---|---|---|---|---|---|
| N | 3428 hCoV-19/Scotland/QEUH-9F7DEE/2020\|EPI_ISL_600917\|2 | ···················· | ···················· | ···················· | ···················· | ···················· | ···················· |
| N | 3429 hCoV-19/Scotland/QEUH-9F9502/2020\|EPI_ISL_600920\|2 | ···················· | ···················· | ···················· | ···················· | ···················· | ···················· |
| N | 3430 hCoV-19/Scotland/QEUH-9F949C/2020\|EPI_ISL_600922\|2 | ···················· | ···················· | ···················· | ···················· | ···················· | ···················· |
| N | 3431 hCoV-19/Scotland/QEUH-9F7DC0/2020\|EPI_ISL_600924\|2 | ···················· | ···················· | ···················· | ···················· | ···················· | ···················· |
| N | 3432 hCoV-19/Wales/MILK-9EF52D/2020\|EPI_ISL_600925\|2020 | ···················· | ···················· | ···················· | ···················· | ···················· | ···················· |
| N | 3433 hCoV-19/Wales/MILK-9F1A29/2020\|EPI_ISL_600926\|2020 | ···················· | ···················· | ···················· | ···················· | ···················· | ···················· |
| N | 3434 hCoV-19/Wales/MILK-9F19FF/2020\|EPI_ISL_600927\|2020 | ···················· | ···················· | ···················· | ···················· | ···················· | ···················· |
| N | 3435 hCoV-19/Wales/MILK-9F16AD/2020\|EPI_ISL_600928\|2020 | ···················· | ···················· | ···················· | ···················· | ···················· | ···················· |
| N | 3436 hCoV-19/Wales/MILK-9EF53C/2020\|EPI_ISL_600929\|2020 | ···················· | ···················· | ···················· | ···················· | ···················· | ···················· |
| N | 3437 hCoV-19/Wales/MILK-9F1A1A/2020\|EPI_ISL_600930\|2020 | ···················· | ···················· | ···················· | ···················· | ···················· | ···················· |
| N | 3438 hCoV-19/England/MILK-9F12B8/2020\|EPI_ISL_600931\|20 | ···················· | ·A·················· | ···················· | ···················· | ···················· | ···················· |
| N | 3439 hCoV-19/Scotland/QEUH-9E5615/2020\|EPI_ISL_600932\|2 | ···················· | ···················· | ···················· | ···················· | ···················· | ···················· |
| N | 3440 hCoV-19/Scotland/QEUH-9E55BE/2020\|EPI_ISL_600933\|2 | ···················· | ···················· | ···················· | ···················· | ···················· | ···················· |
| N | 3441 hCoV-19/Scotland/QEUH-9E4889/2020\|EPI_ISL_600934\|2 | ···················· | ···················· | ···················· | ···················· | ···················· | ···················· |
| N | 3442 hCoV-19/Scotland/QEUH-9E44EF/2020\|EPI_ISL_600935\|2 | ···················· | ···················· | ···················· | ···················· | ···················· | ···················· |
| N | 3443 hCoV-19/Scotland/QEUH-9E4A47/2020\|EPI_ISL_600936\|2 | ···················· | ···················· | ···················· | ···················· | ···················· | ···················· |
| N | 3444 hCoV-19/Scotland/QEUH-9E4D7B/2020\|EPI_ISL_600937\|2 | ···················· | ···················· | ···················· | ···················· | ···················· | ···················· |
| N | 3445 hCoV-19/Scotland/QEUH-9E4D8A/2020\|EPI_ISL_600938\|2 | ···················· | ···················· | ···················· | ···················· | ···················· | ···················· |
| N | 3446 hCoV-19/Scotland/QEUH-9E4300/2020\|EPI_ISL_600939\|2 | ···················· | ···················· | ···················· | ···················· | ···················· | ···················· |
| N | 3447 hCoV-19/Scotland/QEUH-9E534B/2020\|EPI_ISL_600940\|2 | ···················· | ···················· | ···················· | ···················· | ···················· | ···················· |
| N | 3448 hCoV-19/Scotland/QEUH-9E531E/2020\|EPI_ISL_600941\|2 | ···················· | ···················· | ···················· | ···················· | ···················· | ···················· |
| N | 3449 hCoV-19/Scotland/QEUH-9E4634/2020\|EPI_ISL_600942\|2 | ···················· | ···················· | ···················· | ···················· | ···················· | ···················· |
| N | 3450 hCoV-19/Scotland/QEUH-9E5211/2020\|EPI_ISL_600943\|2 | ···················· | ···················· | ···················· | ···················· | ···················· | ···················· |
| N | 3451 hCoV-19/Scotland/QEUH-9E48B6/2020\|EPI_ISL_600944\|2 | ···················· | ···················· | ···················· | ···················· | ···················· | ···················· |
| N | 3452 hCoV-19/Scotland/QEUH-9E4F0C/2020\|EPI_ISL_600945\|2 | ···················· | ···················· | ···················· | ···················· | ···················· | ···················· |
| N | 3453 hCoV-19/Scotland/QEUH-9E46AD/2020\|EPI_ISL_600946\|2 | ···················· | ···················· | ···················· | ···················· | ···················· | ···················· |
| N | 3454 hCoV-19/Scotland/QEUH-9E533C/2020\|EPI_ISL_600947\|2 | ···················· | ···················· | ···················· | ···················· | ···················· | ···················· |
| N | 3455 hCoV-19/Scotland/QEUH-9E4133/2020\|EPI_ISL_600948\|2 | ···················· | ···················· | ···················· | ···················· | ····T··············· | ···················· |
| N | 3456 hCoV-19/Scotland/QEUH-9E51C9/2020\|EPI_ISL_600949\|2 | ···················· | ···················· | ···················· | ···················· | ···················· | ···················· |
| N | 3457 hCoV-19/Scotland/QEUH-9E4898/2020\|EPI_ISL_600950\|2 | ···················· | ···················· | ···················· | ···················· | ···················· | ···················· |
| N | 3458 hCoV-19/Scotland/QEUH-9E5572/2020\|EPI_ISL_600951\|2 | ···················· | ···················· | ···················· | ···················· | ···················· | ···················· |
| N | 3459 hCoV-19/Scotland/QEUH-9E5114/2020\|EPI_ISL_600952\|2 | ···················· | ···················· | ···················· | ···················· | ···················· | ···················· |
| N | 3460 hCoV-19/Scotland/QEUH-9E3ED3/2020\|EPI_ISL_600953\|2 | ···················· | ···················· | ···················· | ···················· | ···················· | ···················· |
| N | 3461 hCoV-19/Scotland/QEUH-9E5220/2020\|EPI_ISL_600998\|2 | ···················· | ···················· | ···················· | ···················· | ···················· | ···················· |
| N | 3462 hCoV-19/England/QEUH-9E53C3/2020\|EPI_ISL_600999\|20 | ···················· | ···················· | ···················· | ···················· | ····T··············· | ···················· |
| N | 3463 hCoV-19/Lithuania/MR-LUHS-Eilnr73/2020\|EPI_ISL_603 | ···················· | ···················· | ···················· | ···················· | ···················· | ···················· |
| N | 3464 hCoV-19/Lithuania/MR-LUHS-Eilnr75/2020\|EPI_ISL_603 | ···················· | ···················· | ···················· | ···················· | ···················· | ···················· |
| N | 3465 hCoV-19/Lithuania/MR-LUHS-Eilnr76/2020\|EPI_ISL_603 | ···················· | ···················· | ···················· | ···················· | ···················· | ···················· |
| N | 3466 hCoV-19/Lithuania/MR-LUHS-Eilnr83/2020\|EPI_ISL_603 | ···················· | ···················· | ···················· | ···················· | ···················· | ···················· |
| N | 3467 hCoV-19/Lithuania/MR-LUHS-Eilnr86/2020\|EPI_ISL_603 | ···················· | ···················· | ···················· | ···················· | ···················· | ···················· |
| N | 3468 hCoV-19/Lithuania/MR-LUHS-Eilnr88/2020\|EPI_ISL_603 | ···················· | ···················· | ···················· | ···················· | ···················· | ···················· |
| N | 3469 hCoV-19/Lithuania/MR-LUHS-Eilnr91/2020\|EPI_ISL_603 | ···················· | ···················· | ···················· | ···················· | ···················· | ···················· |
| N | 3470 hCoV-19/Lithuania/MR-LUHS-Eilnr92/2020\|EPI_ISL_603 | ···················· | ···················· | ···················· | ···················· | ···················· | ···················· |
| N | 3471 hCoV-19/Lithuania/MR-LUHS-Eilnr93/2020\|EPI_ISL_603 | ···················· | ···················· | ···················· | ···················· | ···················· | ···················· |
| N | 3472 hCoV-19/Lithuania/MR-LUHS-Eilnr98/2020\|EPI_ISL_603 | ···················· | ···················· | ···················· | ···················· | ···················· | ···················· |
| N | 3473 hCoV-19/England/204191912/2020\|EPI_ISL_603193\|2020 | ···················· | ···················· | ···················· | ···················· | ···················· | ···················· |
| N | 3474 hCoV-19/England/204191913/2020\|EPI_ISL_603194\|2020 | ···················· | ···················· | ···················· | ···················· | ···················· | ···················· |
| N | 3475 hCoV-19/England/204191995/2020\|EPI_ISL_603195\|2020 | ···················· | ···················· | ···················· | ···················· | ···················· | ···················· |
| N | 3476 hCoV-19/England/204191997/2020\|EPI_ISL_603196\|2020 | ···················· | ···················· | ···················· | ···················· | ···················· | ···················· |
| N | 3477 hCoV-19/England/204191999/2020\|EPI_ISL_603197\|2020 | ···················· | ···················· | ···················· | ···················· | ···················· | ···················· |
| N | 3478 hCoV-19/England/204192000/2020\|EPI_ISL_603198\|2020 | ···················· | ···················· | ···················· | ···················· | ···················· | ···················· |
| N | 3479 hCoV-19/England/204201032/2020\|EPI_ISL_603199\|2020 | ···················· | ···················· | ···················· | ···················· | ···················· | ···················· |
| N | 3480 hCoV-19/USA/CA-QDX-2085/2020\|EPI_ISL_604400\|2020-0 | ···················· | ···················· | ···················· | ···················· | ···················· | ···················· |
| N | 3481 hCoV-19/USA/CA-QDX-2088/2020\|EPI_ISL_604401\|2020-0 | ···················· | ···················· | ···················· | ···················· | ···················· | ···················· |
| N | 3482 hCoV-19/USA/CA-QDX-2089/2020\|EPI_ISL_604402\|2020-0 | ···················· | ···················· | ···················· | ···················· | ···················· | ···················· |
| N | 3483 hCoV-19/USA/CA-QDX-2091/2020\|EPI_ISL_604403\|2020-0 | ···················· | ···················· | ···················· | ···················· | ···················· | ···················· |
| N | 3484 hCoV-19/USA/CA-QDX-2092/2020\|EPI_ISL_604404\|2020-0 | ···················· | ···················· | ···················· | ···················· | ···················· | ···················· |
| N | 3485 hCoV-19/USA/CA-QDX-2093/2020\|EPI_ISL_604405\|2020-0 | ···················· | ···················· | ···················· | ···················· | ···················· | ···················· |
| N | 3486 hCoV-19/USA/CA-QDX-2096/2020\|EPI_ISL_604406\|2020-0 | ···················· | ···················· | ···················· | ···················· | ···················· | ···················· |
| N | 3487 hCoV-19/USA/CA-QDX-2097/2020\|EPI_ISL_604407\|2020-0 | ···················· | ···················· | ···················· | ···················· | ···················· | ···················· |
| N | 3488 hCoV-19/USA/CA-QDX-2098/2020\|EPI_ISL_604408\|2020-0 | ···················· | ···················· | ···················· | ···················· | ···················· | ···················· |
| N | 3489 hCoV-19/USA/CA-QDX-2099/2020\|EPI_ISL_604409\|2020-0 | ···················· | ···················· | ···················· | ···················· | ···················· | ···················· |
| N | 3490 hCoV-19/USA/CA-QDX-2101/2020\|EPI_ISL_604410\|2020-0 | ···················· | ···················· | ···················· | ···················· | ···················· | ···················· |
| N | 3491 hCoV-19/USA/CA-QDX-2102/2020\|EPI_ISL_604411\|2020-0 | ···················· | ···················· | ···················· | ···················· | ···················· | ···················· |
| N | 3492 hCoV-19/USA/CA-QDX-2105/2020\|EPI_ISL_604412\|2020-0 | ···················· | ···················· | ···················· | ···················· | ···················· | ···················· |
| N | 3493 hCoV-19/USA/CA-QDX-2107/2020\|EPI_ISL_604413\|2020-0 | ···················· | ···················· | ···················· | ···················· | ···················· | ···················· |
| N | 3494 hCoV-19/USA/CA-QDX-2109/2020\|EPI_ISL_604414\|2020-0 | ···················· | ···················· | ···················· | ···················· | ···················· | ···················· |
| N | 3495 hCoV-19/USA/CA-QDX-2113/2020\|EPI_ISL_604415\|2020-0 | ···················· | ···················· | ···················· | ···················· | ···················· | ···················· |
| N | 3496 hCoV-19/USA/CA-QDX-2114/2020\|EPI_ISL_604416\|2020-0 | ···················· | ···················· | ···················· | ···················· | ···················· | ···················· |
| N | 3497 hCoV-19/USA/CA-QDX-2116/2020\|EPI_ISL_604417\|2020-0 | ···················· | ···················· | ···················· | ···················· | ···················· | ···················· |
| N | 3498 hCoV-19/USA/CA-QDX-2120/2020\|EPI_ISL_604418\|2020-0 | ···················· | ···················· | ···················· | ···················· | ···················· | ···················· |
| N | 3499 hCoV-19/USA/CA-QDX-2124/2020\|EPI_ISL_604419\|2020-0 | ···················· | ···················· | ···················· | ···················· | ···················· | ···················· |
| N | 3500 hCoV-19/USA/CA-QDX-2127/2020\|EPI_ISL_604420\|2020-0 | ···················· | ···················· | ···················· | ···················· | ···················· | ···················· |
| N | 3501 hCoV-19/USA/CA-QDX-2130/2020\|EPI_ISL_604421\|2020-0 | ···················· | ···················· | ···················· | ···················· | ···················· | ···················· |
| N | 3502 hCoV-19/USA/CA-QDX-2133/2020\|EPI_ISL_604422\|2020-0 | ···················· | ···················· | ···················· | ···················· | ···················· | ···················· |
| N | 3503 hCoV-19/USA/CA-QDX-2134/2020\|EPI_ISL_604423\|2020-0 | ···················· | ···················· | ···················· | ···················· | ···················· | ···················· |
| N | 3504 hCoV-19/USA/CA-QDX-2136/2020\|EPI_ISL_604424\|2020-0 | ···················· | ···················· | ···················· | ···················· | ···················· | ···················· |
| N | 3505 hCoV-19/USA/CA-QDX-2137/2020\|EPI_ISL_604425\|2020-0 | ···················· | ···················· | ···················· | ···················· | ···················· | ···················· |
| N | 3506 hCoV-19/USA/CA-QDX-2138/2020\|EPI_ISL_604426\|2020-0 | ···················· | ···················· | ···················· | ···················· | ···················· | ···················· |
| N | 3507 hCoV-19/USA/CA-QDX-2142/2020\|EPI_ISL_604427\|2020-0 | ···················· | ···················· | ···················· | ···················· | ···················· | ···················· |

| | | | | | | | | |
|---|---|---|---|---|---|---|---|---|
| N | 3508 | hCoV-19/USA/CA-QDX-2143/2020\|EPI_ISL_604428\|2020-0 | ···················· | ···················· | ···················· | ···················· | ···················· | ···················· |
| N | 3509 | hCoV-19/USA/CA-QDX-2144/2020\|EPI_ISL_604429\|2020-0 | ···················· | ···················· | ···················· | ···················· | ···················· | ···················· |
| N | 3510 | hCoV-19/USA/CA-QDX-2148/2020\|EPI_ISL_604430\|2020-0 | ···················· | ···················· | ···················· | ···················· | ···················· | ···················· |
| N | 3511 | hCoV-19/USA/CA-QDX-2152/2020\|EPI_ISL_604431\|2020-0 | ···················· | ···················· | ···················· | ···················· | ···················· | ···················· |
| N | 3512 | hCoV-19/USA/CA-QDX-2155/2020\|EPI_ISL_604432\|2020-0 | ···················· | ···················· | ···················· | ···················· | ···················· | ···················· |
| N | 3513 | hCoV-19/USA/CA-QDX-2157/2020\|EPI_ISL_604433\|2020-0 | ···················· | ···················· | ···················· | ···················· | ···················· | ···················· |
| N | 3514 | hCoV-19/USA/CA-QDX-2159/2020\|EPI_ISL_604434\|2020-0 | ···················· | ···················· | ···················· | ···················· | ···················· | ···················· |
| N | 3515 | hCoV-19/USA/CA-QDX-2160/2020\|EPI_ISL_604435\|2020-0 | ···················· | ···················· | ···················· | ···················· | ···················· | ···················· |
| N | 3516 | hCoV-19/USA/CA-QDX-2162/2020\|EPI_ISL_604436\|2020-0 | ···················· | ···················· | ···················· | ···················· | ···················· | ···················· |
| N | 3517 | hCoV-19/USA/CA-QDX-2166/2020\|EPI_ISL_604437\|2020-0 | ···················· | ···················· | ···················· | ···················· | ···················· | ···················· |
| N | 3518 | hCoV-19/USA/CA-QDX-2169/2020\|EPI_ISL_604438\|2020-0 | ···················· | ···················· | ···················· | ···················· | ···················· | ···················· |
| N | 3520 | hCoV-19/USA/WI-UW-1829/2020\|EPI_ISL_605701\|2020-09 | ···················· | ···················· | ···················· | ···················· | ···················· | ···················· |
| N | 3521 | hCoV-19/USA/WI-UW-1830/2020\|EPI_ISL_605702\|2020-09 | ···················· | ···················· | ···················· | ···················· | ···················· | ···················· |
| N | 3523 | hCoV-19/USA/WI-UW-1832/2020\|EPI_ISL_605704\|2020-09 | ···················· | ···················· | ···················· | ···················· | ···················· | ···················· |
| N | 3524 | hCoV-19/USA/WI-UW-1833/2020\|EPI_ISL_605705\|2020-09 | ···················· | ···················· | ···················· | ···················· | ···················· | ···················· |
| N | 3525 | hCoV-19/USA/WI-UW-1835/2020\|EPI_ISL_605707\|2020-09 | ···················· | ···················· | ···················· | ···················· | ···················· | ···················· |
| N | 3526 | hCoV-19/USA/WI-UW-1836/2020\|EPI_ISL_605708\|2020-09 | ···················· | ···················· | ···················· | ···················· | ···················· | ···················· |
| N | 3527 | hCoV-19/USA/WI-UW-1837/2020\|EPI_ISL_605709\|2020-09 | ···················· | ···················· | ···················· | ···················· | ···················· | ···················· |
| N | 3530 | hCoV-19/USA/WI-UW-1840/2020\|EPI_ISL_605712\|2020-09 | ···················· | ···················· | ···················· | ···················· | ···················· | ···················· |
| N | 3531 | hCoV-19/USA/WI-UW-1841/2020\|EPI_ISL_605713\|2020-09 | ···················· | ···················· | ···················· | ···················· | ···················· | ···················· |
| N | 3532 | hCoV-19/USA/WI-UW-1842/2020\|EPI_ISL_605714\|2020-09 | ···················· | ···················· | ···················· | ···················· | ···················· | ···················· |
| N | 3533 | hCoV-19/USA/WI-UW-1843/2020\|EPI_ISL_605715\|2020-09 | ···················· | ···················· | ···················· | ···················· | ···················· | ···················· |
| N | 3534 | hCoV-19/USA/WI-UW-1844/2020\|EPI_ISL_605716\|2020-09 | ···················· | ···················· | ···················· | ···················· | ···················· | ···················· |
| N | 3535 | hCoV-19/USA/WI-UW-1845/2020\|EPI_ISL_605717\|2020-09 | ···················· | ···················· | ···················· | ···················· | ···················· | ···················· |
| N | 3536 | hCoV-19/USA/WI-UW-1846/2020\|EPI_ISL_605718\|2020-09 | ···················· | ···················· | ···················· | ···················· | ···················· | ···················· |
| N | 3537 | hCoV-19/USA/WI-UW-1847/2020\|EPI_ISL_605719\|2020-09 | ···················· | ···················· | ···················· | ···················· | ···················· | ···················· |
| N | 3539 | hCoV-19/USA/WI-UW-1849/2020\|EPI_ISL_605721\|2020-09 | ···················· | ···················· | ···················· | ···················· | ···················· | ···················· |
| N | 3544 | hCoV-19/USA/WI-UW-1854/2020\|EPI_ISL_605726\|2020-09 | ···················· | ·············A······ | ···················· | ···················· | ···················· | ···················· |
| N | 3549 | hCoV-19/USA/WI-UW-1859/2020\|EPI_ISL_605731\|2020-09 | ···················· | ···················· | ···················· | ···················· | ···················· | ···················· |
| N | 3553 | hCoV-19/USA/WI-UW-1863/2020\|EPI_ISL_605735\|2020-09 | ···················· | ···················· | ···················· | ···················· | ···················· | ···················· |
| N | 3554 | hCoV-19/USA/WI-UW-1864/2020\|EPI_ISL_605736\|2020-09 | ···················· | ···················· | ···················· | ···················· | ···················· | ···················· |
| N | 3562 | hCoV-19/USA/WI-UW-1873/2020\|EPI_ISL_605745\|2020-09 | ···················· | ···················· | ···················· | ···················· | ···················· | ···················· |
| N | 3563 | hCoV-19/USA/WI-UW-1874/2020\|EPI_ISL_605746\|2020-09 | ···················· | ···················· | ···················· | ···················· | ···················· | ···················· |
| N | 3565 | hCoV-19/USA/WI-UW-1876/2020\|EPI_ISL_605748\|2020-09 | ···················· | ···················· | ···················· | ···················· | ···················· | ···················· |
| N | 3566 | hCoV-19/USA/WI-UW-1878/2020\|EPI_ISL_605749\|2020-09 | ···················· | ···················· | ···················· | ···················· | ···················· | ···················· |
| N | 3568 | hCoV-19/USA/WI-UW-1880/2020\|EPI_ISL_605751\|2020-09 | ···················· | ···················· | ···················· | ···················· | ···················· | ···················· |
| N | 3571 | hCoV-19/USA/WI-UW-1886/2020\|EPI_ISL_605755\|2020-09 | ···················· | ···················· | ···················· | ···················· | ···················· | ···················· |
| N | 3572 | hCoV-19/USA/WI-UW-1887/2020\|EPI_ISL_605756\|2020-09 | ···················· | ···················· | ···················· | ···················· | ···················· | ···················· |
| N | 3573 | hCoV-19/USA/WI-UW-1888/2020\|EPI_ISL_605757\|2020-09 | ···················· | ···················· | ···················· | ···················· | ···················· | ···················· |
| N | 3574 | hCoV-19/USA/WI-UW-1889/2020\|EPI_ISL_605758\|2020-09 | ···················· | ···················· | ···················· | ···················· | ···················· | ···················· |
| N | 3575 | hCoV-19/USA/WI-UW-1890/2020\|EPI_ISL_605759\|2020-09 | ···················· | ···················· | ···················· | ···················· | ···················· | ···················· |
| N | 3576 | hCoV-19/USA/WI-UW-1891/2020\|EPI_ISL_605760\|2020-09 | ···················· | ···················· | ···················· | ···················· | ···················· | ···················· |
| N | 3577 | hCoV-19/USA/WI-UW-1892/2020\|EPI_ISL_605761\|2020-09 | ···················· | ···················· | ···················· | ···················· | ···················· | ···················· |
| N | 3579 | hCoV-19/USA/WI-UW-1894/2020\|EPI_ISL_605763\|2020-09 | ···················· | ···················· | ···················· | ···················· | ···················· | ···················· |
| N | 3580 | hCoV-19/USA/WI-UW-1895/2020\|EPI_ISL_605764\|2020-09 | ···················· | ···················· | ···················· | ···················· | ···················· | ···················· |
| N | 3581 | hCoV-19/USA/WI-UW-1896/2020\|EPI_ISL_605765\|2020-09 | ···················· | ···················· | ···················· | ···················· | ···················· | ···················· |
| N | 3582 | hCoV-19/USA/WI-UW-1897/2020\|EPI_ISL_605766\|2020-09 | ···················· | ···················· | ···················· | ···················· | ···················· | ···················· |
| N | 3583 | hCoV-19/USA/WI-UW-1899/2020\|EPI_ISL_605768\|2020-06 | ···················· | ···················· | ···················· | ···················· | ···················· | ···················· |
| N | 3584 | hCoV-19/USA/WI-UW-1900/2020\|EPI_ISL_605769\|2020-09 | ···················· | ···················· | ···················· | ···················· | ···················· | ···················· |
| N | 3585 | hCoV-19/England/ALDP-A520BA/2020\|EPI_ISL_598006\|20 | ···················· | ···················· | ···················· | ···················· | ······T············· | ···················· |
| N | 3586 | hCoV-19/England/ALDP-A51358/2020\|EPI_ISL_598007\|20 | ···················· | ···················· | ···················· | ···················· | ···················· | ···················· |
| N | 3587 | hCoV-19/England/ALDP-A525BB/2020\|EPI_ISL_598008\|20 | ···················· | ···················· | ···················· | ···················· | ···················· | ···················· |
| N | 3588 | hCoV-19/England/ALDP-A52296/2020\|EPI_ISL_598009\|20 | ···················· | ···················· | ···················· | ···················· | ···················· | ···················· |
| N | 3589 | hCoV-19/England/ALDP-A51473/2020\|EPI_ISL_598010\|20 | ···················· | ···················· | ···················· | ···················· | ···················· | ···················· |
| N | 3590 | hCoV-19/England/ALDP-A51622/2020\|EPI_ISL_598011\|20 | ···················· | ···················· | ···················· | ···················· | ···················· | ···················· |
| N | 3591 | hCoV-19/England/ALDP-A52849/2020\|EPI_ISL_598012\|20 | ···················· | ···················· | ···················· | ···················· | ···················· | ···················· |
| N | 3592 | hCoV-19/England/ALDP-A5259D/2020\|EPI_ISL_598013\|20 | ···················· | ···················· | ···················· | ···················· | ···················· | ···················· |
| N | 3593 | hCoV-19/England/ALDP-A51B41/2020\|EPI_ISL_598014\|20 | ···················· | ···················· | ···················· | ···················· | ···················· | ···················· |
| N | 3594 | hCoV-19/England/ALDP-A51E2A/2020\|EPI_ISL_598015\|20 | ···················· | ···················· | ···················· | ···················· | ···················· | ···················· |
| N | 3595 | hCoV-19/England/ALDP-A51446/2020\|EPI_ISL_598017\|20 | ···················· | ···················· | ···················· | ···················· | ···················· | ···················· |
| N | 3596 | hCoV-19/England/ALDP-A519ED/2020\|EPI_ISL_598019\|20 | ···················· | ···················· | ···················· | ···················· | ···················· | ···················· |
| N | 3597 | hCoV-19/England/ALDP-A5158F/2020\|EPI_ISL_598020\|20 | ···················· | ···················· | ···················· | ···················· | ···················· | ···················· |
| N | 3598 | hCoV-19/England/ALDP-A520E7/2020\|EPI_ISL_598021\|20 | ···················· | ···················· | ···················· | ···················· | ···················· | ···················· |
| N | 3599 | hCoV-19/England/ALDP-A516C8/2020\|EPI_ISL_598022\|20 | ···················· | ···················· | ···················· | ···················· | ···················· | ···················· |
| N | 3600 | hCoV-19/England/ALDP-A519A1/2020\|EPI_ISL_598023\|20 | ···················· | ···················· | ···················· | ···················· | ···················· | ···················· |
| N | 3601 | hCoV-19/England/ALDP-A51A35/2020\|EPI_ISL_598024\|20 | ···················· | ···················· | ···················· | ···················· | ···················· | ···················· |
| N | 3602 | hCoV-19/England/ALDP-A51613/2020\|EPI_ISL_598025\|20 | ···················· | ···················· | ···················· | ···················· | ···················· | ···················· |
| N | 3603 | hCoV-19/England/ALDP-A51543/2020\|EPI_ISL_598026\|20 | ···················· | ···················· | ···················· | ···················· | ···················· | ···················· |
| N | 3604 | hCoV-19/England/ALDP-A51BC9/2020\|EPI_ISL_598027\|20 | ···················· | ···················· | ···················· | ···················· | ···················· | ···················· |
| N | 3605 | hCoV-19/England/ALDP-A51D1E/2020\|EPI_ISL_598028\|20 | ···················· | ···················· | ···················· | ···················· | ···················· | ···················· |
| N | 3606 | hCoV-19/England/ALDP-A52463/2020\|EPI_ISL_598029\|20 | ···················· | ···················· | ···················· | ···················· | ···················· | ···················· |
| N | 3607 | hCoV-19/England/ALDP-A5267C/2020\|EPI_ISL_598030\|20 | ···················· | ···················· | ···················· | ···················· | ···················· | ···················· |
| N | 3608 | hCoV-19/England/ALDP-A51877/2020\|EPI_ISL_598039\|20 | ···················· | ···················· | ···················· | ···················· | ···················· | ···················· |
| N | 3609 | hCoV-19/England/ALDP-A5174D/2020\|EPI_ISL_598040\|20 | ···················· | ···················· | ···················· | ···················· | ···················· | ···················· |
| N | 3610 | hCoV-19/England/ALDP-A51CB7/2020\|EPI_ISL_598041\|20 | ···················· | ···················· | ···················· | ···················· | ···················· | ···················· |
| N | 3611 | hCoV-19/England/ALDP-A526D6/2020\|EPI_ISL_598042\|20 | ···················· | ···················· | ···················· | ···················· | ···················· | ···················· |
| N | 3612 | hCoV-19/England/ALDP-A514A0/2020\|EPI_ISL_598043\|20 | ···················· | ···················· | ···················· | ···················· | ···················· | ···················· |
| N | 3613 | hCoV-19/England/ALDP-A512C4/2020\|EPI_ISL_598044\|20 | ···················· | ···················· | ···················· | ···················· | ···················· | ···················· |
| N | 3614 | hCoV-19/England/ALDP-A51A80/2020\|EPI_ISL_598045\|20 | ···················· | ···················· | ···················· | ···················· | ···················· | ···················· |
| N | 3615 | hCoV-19/England/ALDP-A511F4/2020\|EPI_ISL_598046\|20 | ···················· | ···················· | ···················· | ···················· | ···················· | ···················· |

N 3616 hCoV-19/England/ALDP-A51ABD/2020|EPI_ISL_598047|20 ···················· ···················· ···················· ···················· ···················· ····················
N 3617 hCoV-19/England/ALDP-A528EF/2020|EPI_ISL_598048|20 ···················· ···················· ···················· ···················· ···················· ····················
N 3618 hCoV-19/England/ALDP-A5272E/2020|EPI_ISL_598049|20 ···················· ···················· ···················· ···················· ···················· ····················
N 3619 hCoV-19/England/ALDP-A5269A/2020|EPI_ISL_598052|20 ···················· ···················· ···················· ···················· ···················· ····················
N 3620 hCoV-19/England/ALDP-A51CA8/2020|EPI_ISL_598053|20 ···················· ···················· ···················· ···················· ···················· ····················
N 3621 hCoV-19/England/ALDP-A515DA/2020|EPI_ISL_598054|20 ···················· ···················· ···················· ···················· ···················· ····················
N 3622 hCoV-19/England/ALDP-A52876/2020|EPI_ISL_598055|20 ···················· ···················· ···················· ···················· ···················· ····················
N 3623 hCoV-19/England/ALDP-A51DB4/2020|EPI_ISL_598056|20 ···················· ···················· ···················· ···················· ···················· ····················
N 3624 hCoV-19/England/ALDP-A52515/2020|EPI_ISL_598057|20 ···················· ···················· ···················· ···················· ···················· ····················
N 3625 hCoV-19/England/ALDP-A51C3F/2020|EPI_ISL_598058|20 ···················· ···················· ···················· ···················· ···················· ····················
N 3626 hCoV-19/England/ALDP-A5221E/2020|EPI_ISL_598059|20 ···················· ···················· ···················· ···················· ···················· ····················
N 3627 hCoV-19/England/ALDP-A51376/2020|EPI_ISL_598060|20 ···················· ·A·················· ···················· ···················· ···················· ····················
N 3628 hCoV-19/England/ALDP-A51798/2020|EPI_ISL_598061|20 ···················· ···················· ···················· ···················· ···················· ····················
N 3629 hCoV-19/England/ALDP-A528FE/2020|EPI_ISL_598062|20 ···················· ···················· ···················· ···················· ···················· ····················
N 3630 hCoV-19/England/ALDP-A52269/2020|EPI_ISL_598063|20 ···················· ·A·················· ···················· ···················· ···················· ····················
N 3631 hCoV-19/England/ALDP-A52278/2020|EPI_ISL_598065|20 ···················· ···················· ···················· ···················· ···················· ····················
N 3632 hCoV-19/England/ALDP-A515CB/2020|EPI_ISL_598066|20 ···················· ···················· ···················· ···················· ···················· ····················
N 3633 hCoV-19/England/ALDP-A517B6/2020|EPI_ISL_598067|20 ···················· ···················· ···················· ···················· ···················· ····················
N 3634 hCoV-19/England/ALDP-A52797/2020|EPI_ISL_598068|20 ···················· ···················· ···················· ···················· ···················· ····················
N 3635 hCoV-19/England/ALDP-A51EFD/2020|EPI_ISL_598069|20 ···················· ···················· ···················· ···················· ···················· ····················
N 3636 hCoV-19/England/ALDP-A5159E/2020|EPI_ISL_598070|20 ···················· ···················· ···················· ···················· ···················· ····················
N 3637 hCoV-19/England/ALDP-A5121F/2020|EPI_ISL_598071|20 ···················· ···················· ···················· ···················· ···················· ····················
N 3638 hCoV-19/England/ALDP-A51C5D/2020|EPI_ISL_598072|20 ···················· ···················· ···················· ···················· ···················· ····················
N 3639 hCoV-19/England/ALDP-A52375/2020|EPI_ISL_598073|20 ···················· ···················· ···················· ···················· ······T············· ····················
N 3640 hCoV-19/England/ALDP-A5280D/2020|EPI_ISL_598074|20 ···················· ···················· ···················· ···················· ···················· ····················
N 3641 hCoV-19/England/ALDP-A52111/2020|EPI_ISL_598076|20 ···················· ···················· ···················· ···················· ···················· ····················
N 3642 hCoV-19/England/ALDP-A515F8/2020|EPI_ISL_598077|20 ···················· ···················· ···················· ···················· ···················· ····················
N 3643 hCoV-19/England/ALDP-A51D2D/2020|EPI_ISL_598079|20 ···················· ···················· ···················· ···················· ···················· ····················
N 3644 hCoV-19/England/ALDP-A51200/2020|EPI_ISL_598080|20 ···················· ···················· ···················· ···················· ···················· ····················
N 3645 hCoV-19/England/ALDP-A51349/2020|EPI_ISL_598082|20 ···················· ·A·················· ···················· ···················· ···················· ····················
N 3646 hCoV-19/England/ALDP-A51DA5/2020|EPI_ISL_598083|20 ···················· ·A·················· ···················· ···················· ···················· ····················
N 3647 hCoV-19/England/ALDP-A5126A/2020|EPI_ISL_598084|20 ···················· ···················· ···················· ···················· ···················· ····················
N 3648 hCoV-19/England/ALDP-A51F54/2020|EPI_ISL_598085|20 ···················· ···················· ···················· ···················· ···················· ····················
N 3649 hCoV-19/England/ALDP-A52603/2020|EPI_ISL_598086|20 ···················· ···················· ···················· ···················· ···················· ····················
N 3650 hCoV-19/England/ALDP-A52621/2020|EPI_ISL_598087|20 ···················· ···················· ···················· ···················· ···················· ····················
N 3651 hCoV-19/England/ALDP-A51E93/2020|EPI_ISL_598088|20 ···················· ···················· ···················· ···················· ···················· ····················
N 3652 hCoV-19/England/ALDP-A5265E/2020|EPI_ISL_598089|20 ···················· ···················· ···················· ···················· ···················· ····················
N 3653 hCoV-19/England/QEUH-A0B9B5/2020|EPI_ISL_599337|20 ···················· ···················· ···················· ···················· ···················· ····················
N 3654 hCoV-19/England/QEUH-A0B1BD/2020|EPI_ISL_599338|20 ···················· ···················· ···················· ···················· ···················· ····················
N 3655 hCoV-19/England/QEUH-A0ACEA/2020|EPI_ISL_599339|20 ···················· ···················· ···················· ···················· ···················· ····················
N 3656 hCoV-19/England/QEUH-A0AEB7/2020|EPI_ISL_599340|20 ···················· ·A·················· ···················· ···················· ···················· ····················
N 3657 hCoV-19/Scotland/QEUH-A0C310/2020|EPI_ISL_599341|2 ···················· ···················· ···················· ···················· ···················· ····················
N 3658 hCoV-19/Scotland/QEUH-A0BED4/2020|EPI_ISL_599342|2 ···················· ···················· ···················· ···················· ···················· ····················
N 3660 hCoV-19/Scotland/QEUH-A0BC52/2020|EPI_ISL_599344|2 ···················· ···················· ···················· ···················· ···················· ····················
N 3661 hCoV-19/England/QEUH-A0AF1E/2020|EPI_ISL_599346|20 ···················· ···················· ···················· ···················· ···················· ····················
N 3662 hCoV-19/England/QEUH-A0BAB2/2020|EPI_ISL_599348|20 ···················· ···················· ···················· ···················· ···················· ····················
N 3663 hCoV-19/England/QEUH-A0BE2F/2020|EPI_ISL_599350|20 ···················· ···················· ···················· ···················· ···················· ····················
N 3664 hCoV-19/England/QEUH-A0B038/2020|EPI_ISL_599351|20 ···················· ······C············· ···················· ···················· ···················· ····················
N 3665 hCoV-19/England/QEUH-A0B6CD/2020|EPI_ISL_599352|20 ···················· ···················· ···················· ···················· ····T··············· ····················
N 3666 hCoV-19/Scotland/QEUH-A0B88B/2020|EPI_ISL_599353|2 ···················· ···················· ···················· ···················· ···················· ····················
N 3667 hCoV-19/Scotland/QEUH-A0C28C/2020|EPI_ISL_599354|2 ···················· ···················· ···················· ···················· ···················· ····················
N 3668 hCoV-19/England/QEUH-A0B4F0/2020|EPI_ISL_599355|20 ···················· ···················· ···················· ···················· ···················· ····················
N 3669 hCoV-19/England/QEUH-A0AB0B/2020|EPI_ISL_599356|20 ·······C············ ···················· ···················· ···················· ···················· ····················
N 3670 hCoV-19/Scotland/QEUH-A0B42D/2020|EPI_ISL_599357|2 ···················· ···················· ···················· ···················· ···················· ····················
N 3671 hCoV-19/England/QEUH-A0B715/2020|EPI_ISL_599358|20 ···················· ···················· ···················· ···················· ···················· ····················
N 3672 hCoV-19/England/QEUH-A0B2E7/2020|EPI_ISL_599359|20 ···················· ···················· ···················· ···················· ···················· ····················
N 3673 hCoV-19/Scotland/QEUH-A0AE6C/2020|EPI_ISL_599361|2 ···················· ···················· ···················· ···················· ······T············· ···············A····
N 3674 hCoV-19/England/QEUH-A0B047/2020|EPI_ISL_599370|20 ···················· ···················· ···················· ···················· ···················· ····················
N 3675 hCoV-19/England/QEUH-A0BE01/2020|EPI_ISL_599371|20 ···················· ···················· ···················· ···················· ···················· ····················
N 3676 hCoV-19/England/QEUH-A0BEB6/2020|EPI_ISL_599373|20 ···················· ···················· ···················· ···················· ···················· ····················
N 3677 hCoV-19/England/QEUH-A0BA49/2020|EPI_ISL_599374|20 ···················· ···················· ···················· ···················· ···················· ····················
N 3678 hCoV-19/England/QEUH-A0AB47/2020|EPI_ISL_599375|20 ···················· ···················· ···················· ···················· ···················· ····················
N 3679 hCoV-19/Scotland/QEUH-A0B4B4/2020|EPI_ISL_599376|2 ···················· ···················· ···················· ···················· ···················· ····················
N 3680 hCoV-19/Scotland/QEUH-A0AE20/2020|EPI_ISL_599377|2 ···················· ···················· ···················· ···················· ···················· ····················
N 3681 hCoV-19/Scotland/QEUH-A0BCCB/2020|EPI_ISL_599378|2 ···················· ···················· ···················· ···················· ···················· ····················
N 3682 hCoV-19/Scotland/QEUH-A0BD04/2020|EPI_ISL_599379|2 ···················· ···················· ···················· ···················· ···················· ····················
N 3683 hCoV-19/England/QEUH-A0BAC1/2020|EPI_ISL_599380|20 ···················· ···················· ···················· ···················· ···················· ····················
N 3684 hCoV-19/England/QEUH-A0B636/2020|EPI_ISL_599381|20 ···················· ···················· ···················· ···················· ···················· ····················
N 3685 hCoV-19/England/QEUH-A0C0A0/2020|EPI_ISL_599382|20 ···················· ···················· ···················· ···················· ···················· ····················
N 3686 hCoV-19/Scotland/QEUH-A0ABFC/2020|EPI_ISL_599383|2 ···················· ···················· ···················· ···················· ···················· ····················
N 3687 hCoV-19/Scotland/QEUH-A0BC61/2020|EPI_ISL_599384|2 ···················· ···················· ···················· ···················· ···················· ····················
N 3688 hCoV-19/Scotland/QEUH-A0BD9B/2020|EPI_ISL_599385|2 ···················· ···················· ···················· ···················· ···················· ····················
N 3689 hCoV-19/England/QEUH-A0B1AE/2020|EPI_ISL_599387|20 ···················· ···················· ···················· ···················· ···················· ····················
N 3690 hCoV-19/England/QEUH-A0BBCE/2020|EPI_ISL_599388|20 ···················· ···················· ···················· ···················· ···················· ····················
N 3691 hCoV-19/England/QEUH-A0B33F/2020|EPI_ISL_599389|20 ···················· ···················· ···················· ···················· ···················· ····················
N 3692 hCoV-19/Scotland/QEUH-A0AE3F/2020|EPI_ISL_599390|2 ···················· ···················· ···················· ···················· ···················· ····················
N 3693 hCoV-19/Scotland/QEUH-A0AAB3/2020|EPI_ISL_599391|2 ···················· ···················· ···················· ···················· ···················· ····················
N 3694 hCoV-19/England/QEUH-A0C082/2020|EPI_ISL_599392|20 ···················· ···················· ···················· ···················· ···················· ····················
N 3695 hCoV-19/Scotland/QEUH-A0B91F/2020|EPI_ISL_599393|2 ···················· ···················· ···················· ···················· ···················· ····················
N 3696 hCoV-19/Scotland/QEUH-A0BEF2/2020|EPI_ISL_599394|2 ···················· ···················· ···················· ···················· ···················· ····················

| | | | | | | | | |
|---|---|---|---|---|---|---|---|---|
| N | 3697 | hCoV-19/England/QEUH-A0AEE4/2020\|EPI_ISL_599396\|20 | ···················· | ···················· | ···················· | ···················· | ······T············· | ···················· |
| N | 3698 | hCoV-19/England/QEUH-A0AD41/2020\|EPI_ISL_599397\|20 | ···················· | ···················· | ···················· | ···················· | ···················· | ···················· |
| N | 3699 | hCoV-19/England/QEUH-A0C8D5/2020\|EPI_ISL_599398\|20 | ···················· | ···················· | ···················· | ···················· | ···················· | ···················· |
| N | 3700 | hCoV-19/Scotland/QEUH-A0B706/2020\|EPI_ISL_599399\|2 | ···················· | ···················· | ···················· | ···················· | ···················· | ···················· |
| N | 3701 | hCoV-19/Scotland/QEUH-9F7D75/2020\|EPI_ISL_600900\|2 | ···················· | ···················· | ···················· | ···················· | ···················· | ···················· |
| N | 3702 | hCoV-19/Wales/MILK-9F2457/2020\|EPI_ISL_600901\|2020 | ···················· | ···················· | ···················· | ···················· | ···················· | ···················· |
| N | 3703 | hCoV-19/Scotland/QEUH-9F7407/2020\|EPI_ISL_600902\|2 | ···················· | ···················· | ···················· | ···················· | ···················· | ···················· |
| N | 3704 | hCoV-19/Scotland/QEUH-9F7540/2020\|EPI_ISL_600907\|2 | ···················· | ···················· | ···················· | ···················· | ···················· | ···················· |
| N | 3705 | hCoV-19/Scotland/QEUH-9F948D/2020\|EPI_ISL_600908\|2 | ···················· | ···················· | ···················· | ···················· | ······T············· | ···················· |
| N | 3706 | hCoV-19/Scotland/QEUH-9F7DB1/2020\|EPI_ISL_600909\|2 | ···················· | ···················· | ···················· | ···················· | ···················· | ···················· |
| N | 3707 | hCoV-19/Lithuania/MR-LUHS-Eilnr99/2020\|EPI_ISL_603 | ···················· | ···················· | ···················· | ···················· | ···················· | ···················· |
| N | 3708 | hCoV-19/Lithuania/MR-LUHS-Eilnr100/2020\|EPI_ISL_60 | ···················· | ···················· | ···················· | ···················· | ···················· | ···················· |
| N | 3709 | hCoV-19/Lithuania/KU-LUHS-Eilnr102/2020\|EPI_ISL_60 | ···················· | ···················· | ···················· | ···················· | ···················· | ···················· |
| N | 3710 | hCoV-19/Lithuania/MR-LUHS-Eilnr103/2020\|EPI_ISL_60 | ···················· | ···················· | ···················· | ···················· | ···················· | ···················· |
| N | 3711 | hCoV-19/Lithuania/MR-LUHS-Eilnr104/2020\|EPI_ISL_60 | ···················· | ···················· | ···················· | ···················· | ···················· | ···················· |
| N | 3712 | hCoV-19/Lithuania/KU-LUHS-Eilnr107/2020\|EPI_ISL_60 | ···················· | ···················· | ···················· | ···················· | ···················· | ···················· |
| N | 3713 | hCoV-19/Lithuania/KU-LUHS-Eilnr110/2020\|EPI_ISL_60 | ···················· | ···················· | ···················· | ···················· | ···················· | ···················· |
| N | 3714 | hCoV-19/Lithuania/MR-LUHS-Eilnr111/2020\|EPI_ISL_60 | ···················· | ···················· | ···················· | ···················· | ···················· | ···················· |
| N | 3715 | hCoV-19/Lithuania/KU-LUHS-Eilnr116/2020\|EPI_ISL_60 | ···················· | ···················· | ···················· | ···················· | ···················· | ···················· |
| N | 3716 | hCoV-19/Lithuania/KL-LUHS-Eilnr58/2020\|EPI_ISL_603 | ···················· | ···················· | ···················· | ···················· | ···················· | ···················· |
| N | 3717 | hCoV-19/Australia/SAP517/2020\|EPI_ISL_603123\|2020- | ···················· | ···················· | ···················· | ···················· | ···················· | ···················· |
| N | 3718 | hCoV-19/Australia/SAP519/2020\|EPI_ISL_603124\|2020- | ···················· | ·A·················· | ···················· | ···················· | ···················· | ··A················· |
| N | 3719 | hCoV-19/Australia/SAP520/2020\|EPI_ISL_603125\|2020- | ···················· | ···················· | ···················· | ···················· | ···················· | ···················· |
| N | 3720 | hCoV-19/Australia/SAP516/2020\|EPI_ISL_603128\|2020- | ···················· | ·A·················· | ···················· | ···················· | ···················· | ···················· |
| N | 3721 | hCoV-19/Italy/LAZ-INMI-30/2020\|EPI_ISL_603137\|2020 | ···················· | ···················· | ···················· | ···················· | ···················· | ···················· |
| N | 3722 | hCoV-19/Italy/LAZ-INMI-31/2020\|EPI_ISL_603138\|2020 | ···················· | ···················· | ···················· | ···················· | ···················· | ···················· |
| N | 3723 | hCoV-19/Italy/LAZ-INMI-32/2020\|EPI_ISL_603139\|2020 | ···················· | ···················· | ···················· | ···················· | ···················· | ···················· |
| N | 3724 | hCoV-19/Italy/LAZ-INMI-39/2020\|EPI_ISL_603146\|2020 | ···················· | ···················· | ···················· | ···················· | ···················· | ···················· |
| N | 3725 | hCoV-19/Italy/LAZ-INMI-40/2020\|EPI_ISL_603147\|2020 | ···················· | ···················· | ···················· | ···················· | ···················· | ···················· |
| N | 3726 | hCoV-19/Italy/LAZ-INMI-43/2020\|EPI_ISL_603150\|2020 | ···················· | ···················· | ···················· | ···················· | ···················· | ···················· |
| N | 3727 | hCoV-19/Italy/LAZ-INMI-44/2020\|EPI_ISL_603151\|2020 | ···················· | ···················· | ···················· | ···················· | ···················· | ···················· |
| N | 3728 | hCoV-19/Italy/LAZ-INMI-45/2020\|EPI_ISL_603152\|2020 | ···················· | ···················· | ···················· | ···················· | ···················· | ···················· |
| N | 3729 | hCoV-19/Italy/LAZ-INMI-46/2020\|EPI_ISL_603153\|2020 | ···················· | ···················· | ···················· | ···················· | ···················· | ···················· |
| N | 3730 | hCoV-19/Italy/LAZ-INMI-47/2020\|EPI_ISL_603154\|2020 | ···················· | ···················· | ···················· | ···················· | ···················· | ···················· |
| N | 3731 | hCoV-19/Italy/LAZ-INMI-48/2020\|EPI_ISL_603155\|2020 | ···················· | ···················· | ···················· | ···················· | ···················· | ···················· |
| N | 3732 | hCoV-19/Italy/LAZ-INMI-49/2020\|EPI_ISL_603156\|2020 | ···················· | ···················· | ···················· | ···················· | ···················· | ···················· |
| N | 3733 | hCoV-19/Italy/LAZ-INMI-50/2020\|EPI_ISL_603157\|2020 | ···················· | ···················· | ···················· | ···················· | ···················· | ···················· |
| N | 3734 | hCoV-19/Italy/LAZ-INMI-51/2020\|EPI_ISL_603158\|2020 | ···················· | ···················· | ···················· | ···················· | ···················· | ···················· |
| N | 3735 | hCoV-19/Italy/LAZ-INMI-52/2020\|EPI_ISL_603159\|2020 | ···················· | ···················· | ···················· | ···················· | ···················· | ···················· |
| N | 3736 | hCoV-19/Italy/LAZ-INMI-53/2020\|EPI_ISL_603160\|2020 | ···················· | ···················· | ···················· | ···················· | ···················· | ···················· |
| N | 3737 | hCoV-19/Italy/LAZ-INMI-54/2020\|EPI_ISL_603161\|2020 | ···················· | ···················· | ···················· | ···················· | ···················· | ···················· |
| N | 3738 | hCoV-19/Italy/LAZ-INMI-55/2020\|EPI_ISL_603162\|2020 | ···················· | ···················· | ···················· | ···················· | ···················· | ···················· |
| N | 3739 | hCoV-19/Italy/LAZ-INMI-56/2020\|EPI_ISL_603163\|2020 | ···················· | ···················· | ···················· | ···················· | ···················· | ···················· |
| N | 3740 | hCoV-19/Italy/LAZ-INMI-57/2020\|EPI_ISL_603164\|2020 | ···················· | ···················· | ···················· | ···················· | ···················· | ···················· |
| N | 3741 | hCoV-19/Italy/LAZ-INMI-58/2020\|EPI_ISL_603165\|2020 | ···················· | ···················· | ···················· | ···················· | ···················· | ···················· |
| N | 3742 | hCoV-19/Italy/LAZ-INMI-59/2020\|EPI_ISL_603166\|2020 | ···················· | ···················· | ···················· | ···················· | ···················· | ···················· |
| N | 3743 | hCoV-19/Italy/LAZ-INMI-60/2020\|EPI_ISL_603167\|2020 | ···················· | ···················· | ···················· | ···················· | ···················· | ···················· |
| N | 3744 | hCoV-19/Italy/LAZ-INMI-61/2020\|EPI_ISL_603168\|2020 | ···················· | ···················· | ···················· | ···················· | ···················· | ···················· |
| N | 3745 | hCoV-19/Italy/LAZ-INMI-62/2020\|EPI_ISL_603169\|2020 | ···················· | ···················· | ···················· | ···················· | ···················· | ···················· |
| N | 3746 | hCoV-19/Italy/LAZ-INMI-63/2020\|EPI_ISL_603170\|2020 | ···················· | ···················· | ···················· | ···················· | ···················· | ···················· |
| N | 3747 | hCoV-19/Italy/LAZ-INMI-64/2020\|EPI_ISL_603171\|2020 | ···················· | ···················· | ···················· | ···················· | ···················· | ···················· |
| N | 3748 | hCoV-19/Italy/LAZ-INMI-65/2020\|EPI_ISL_603172\|2020 | ···················· | ···················· | ···················· | ···················· | ···················· | ···················· |
| N | 3749 | hCoV-19/Italy/LAZ-INMI-66/2020\|EPI_ISL_603173\|2020 | ···················· | ···················· | ···················· | ···················· | ···················· | ···················· |
| N | 3750 | hCoV-19/Italy/LAZ-INMI-67/2020\|EPI_ISL_603174\|2020 | ···················· | ···················· | ···················· | ···················· | ···················· | ···················· |
| N | 3751 | hCoV-19/Italy/LAZ-INMI-68/2020\|EPI_ISL_603175\|2020 | ···················· | ···················· | ···················· | ···················· | ···················· | ···················· |
| N | 3752 | hCoV-19/Italy/LAZ-INMI-69/2020\|EPI_ISL_603176\|2020 | ···················· | ···················· | ···················· | ···················· | ···················· | ···················· |
| N | 3753 | hCoV-19/Italy/LAZ-INMI-70/2020\|EPI_ISL_603177\|2020 | ···················· | ···················· | ···················· | ···················· | ···················· | ···················· |
| N | 3754 | hCoV-19/Italy/LAZ-INMI-71/2020\|EPI_ISL_603178\|2020 | ···················· | ···················· | ···················· | ···················· | ···················· | ···················· |
| N | 3755 | hCoV-19/Italy/LAZ-INMI-72/2020\|EPI_ISL_603179\|2020 | ···················· | ···················· | ···················· | ···················· | ···················· | ···················· |
| N | 3756 | hCoV-19/Italy/LAZ-INMI-73/2020\|EPI_ISL_603180\|2020 | ···················· | ···················· | ···················· | ···················· | ···················· | ···················· |
| N | 3757 | hCoV-19/Italy/LAZ-INMI-74/2020\|EPI_ISL_603181\|2020 | ···················· | ···················· | ···················· | ···················· | ···················· | ···················· |
| N | 3758 | hCoV-19/Italy/LAZ-INMI-75/2020\|EPI_ISL_603182\|2020 | ···················· | ···················· | ···················· | ···················· | ···················· | ···················· |
| N | 3759 | hCoV-19/Italy/LAZ-INMI-76/2020\|EPI_ISL_603183\|2020 | ···················· | ···················· | ···················· | ···················· | ···················· | ···················· |
| N | 3760 | hCoV-19/Italy/LAZ-INMI-77/2020\|EPI_ISL_603184\|2020 | ···················· | ···················· | ···················· | ···················· | ···················· | ···················· |
| N | 3761 | hCoV-19/Italy/LAZ-INMI-78/2020\|EPI_ISL_603185\|2020 | ···················· | ···················· | ···················· | ···················· | ···················· | ···················· |
| N | 3762 | hCoV-19/Italy/LAZ-INMI-79/2020\|EPI_ISL_603186\|2020 | ···················· | ···················· | ···················· | ···················· | ···················· | ···················· |
| N | 3763 | hCoV-19/Italy/LAZ-INMI-80/2020\|EPI_ISL_603187\|2020 | ···················· | ···················· | ···················· | ···················· | ···················· | ···················· |
| N | 3764 | hCoV-19/England/204191453/2020\|EPI_ISL_603188\|2020 | ···················· | ···················· | ···················· | ···················· | ···················· | ···················· |
| N | 3765 | hCoV-19/England/204191511/2020\|EPI_ISL_603189\|2020 | ···················· | ···················· | ···················· | ···················· | ···················· | ···················· |
| N | 3766 | hCoV-19/USA/CA-QDX-2171/2020\|EPI_ISL_604440\|2020-0 | ···················· | ···················· | ···················· | ···················· | ···················· | ···················· |
| N | 3767 | hCoV-19/USA/CA-QDX-2174/2020\|EPI_ISL_604441\|2020-0 | ···················· | ···················· | ···················· | ···················· | ···················· | ···················· |
| N | 3768 | hCoV-19/USA/CA-QDX-2175/2020\|EPI_ISL_604442\|2020-0 | ···················· | ···················· | ···················· | ···················· | ···················· | ···················· |
| N | 3769 | hCoV-19/USA/CA-QDX-2179/2020\|EPI_ISL_604443\|2020-0 | ···················· | ···················· | ···················· | ···················· | ···················· | ···················· |
| N | 3770 | hCoV-19/USA/CA-QDX-2180/2020\|EPI_ISL_604444\|2020-0 | ···················· | ···················· | ···················· | ···················· | ···················· | ···················· |
| N | 3771 | hCoV-19/USA/CA-QDX-2181/2020\|EPI_ISL_604445\|2020-0 | ···················· | ···················· | ···················· | ···················· | ···················· | ···················· |
| N | 3772 | hCoV-19/USA/CA-QDX-2185/2020\|EPI_ISL_604446\|2020-0 | ···················· | ···················· | ···················· | ···················· | ···················· | ···················· |
| N | 3773 | hCoV-19/USA/CT-QDX-2094/2020\|EPI_ISL_604447\|2020-0 | ···················· | ···················· | ···················· | ···················· | ···················· | ···················· |
| N | 3774 | hCoV-19/USA/FL-QDX-2177/2020\|EPI_ISL_604448\|2020-0 | ···················· | ···················· | ···················· | ···················· | ···················· | ···················· |
| N | 3775 | hCoV-19/USA/GA-QDX-2115/2020\|EPI_ISL_604449\|2020-0 | ···················· | ···················· | ···················· | ···················· | ···················· | ···················· |
| N | 3776 | hCoV-19/USA/GA-QDX-2150/2020\|EPI_ISL_604450\|2020-0 | ···················· | ···················· | ···················· | ···················· | ···················· | ···················· |

| | | | | | | | | |
|---|---|---|---|---|---|---|---|---|
| N | 3777 | hCoV-19/USA/GA-QDX-2158/2020|EPI_ISL_604451|2020-0 | ···················· | ···················· | ····················· | ···················· | ···················· | ···················· |
| N | 3778 | hCoV-19/USA/HI-QDX-2149/2020|EPI_ISL_604452|2020-0 | ···················· | ···················· | ····················· | ···················· | ···················· | ···················· |
| N | 3779 | hCoV-19/USA/LA-QDX-2100/2020|EPI_ISL_604453|2020-0 | ···················· | ···················· | ····················· | ···················· | ···················· | ···················· |
| N | 3780 | hCoV-19/USA/LA-QDX-2108/2020|EPI_ISL_604454|2020-0 | ···················· | ···················· | ····················· | ···················· | ···················· | ···················· |
| N | 3781 | hCoV-19/USA/LA-QDX-2131/2020|EPI_ISL_604455|2020-0 | ···················· | ···················· | ····················· | ···················· | ···················· | ···················· |
| N | 3782 | hCoV-19/USA/MA-QDX-2106/2020|EPI_ISL_604456|2020-0 | ···················· | ···················· | ····················· | ···················· | ···················· | ···················· |
| N | 3783 | hCoV-19/USA/MI-QDX-2073/2020|EPI_ISL_604457|2020-0 | ···················· | ···················· | ····················· | ···················· | ···················· | ···················· |
| N | 3784 | hCoV-19/USA/MI-QDX-2076/2020|EPI_ISL_604458|2020-0 | ···················· | ···················· | ····················· | ···················· | ···················· | ···················· |
| N | 3785 | hCoV-19/USA/MI-QDX-2086/2020|EPI_ISL_604459|2020-0 | ···················· | ···················· | ····················· | ···················· | ···················· | ···················· |
| N | 3786 | hCoV-19/USA/MI-QDX-2087/2020|EPI_ISL_604460|2020-0 | ···················· | ···················· | ····················· | ···················· | ···················· | ···················· |
| N | 3787 | hCoV-19/USA/MI-QDX-2095/2020|EPI_ISL_604461|2020-0 | ···················· | ···················· | ····················· | ···················· | ···················· | ···················· |
| N | 3788 | hCoV-19/USA/MI-QDX-2110/2020|EPI_ISL_604462|2020-0 | ···················· | ···················· | ····················· | ···················· | ···················· | ···················· |
| N | 3789 | hCoV-19/USA/MI-QDX-2119/2020|EPI_ISL_604463|2020-0 | ···················· | ···················· | ····················· | ···················· | ···················· | ···················· |
| N | 3790 | hCoV-19/USA/MI-QDX-2140/2020|EPI_ISL_604464|2020-0 | ···················· | ···················· | ····················· | ···················· | ···················· | ···················· |
| N | 3791 | hCoV-19/USA/MI-QDX-2141/2020|EPI_ISL_604465|2020-0 | ···················· | ···················· | ····················· | ···················· | ···················· | ···················· |
| N | 3792 | hCoV-19/USA/MI-QDX-2145/2020|EPI_ISL_604466|2020-0 | ···················· | ···················· | ····················· | ···················· | ···················· | ···················· |
| N | 3793 | hCoV-19/USA/MI-QDX-2146/2020|EPI_ISL_604467|2020-0 | ···················· | ···················· | ····················· | ···················· | ···················· | ···················· |
| N | 3794 | hCoV-19/USA/MI-QDX-2147/2020|EPI_ISL_604468|2020-0 | ···················· | ···················· | ····················· | ···················· | ···················· | ···················· |
| N | 3795 | hCoV-19/USA/MI-QDX-2153/2020|EPI_ISL_604469|2020-0 | ···················· | ···················· | ····················· | ···················· | ···················· | ···················· |
| N | 3796 | hCoV-19/USA/MI-QDX-2184/2020|EPI_ISL_604477|2020-0 | ···················· | ···················· | ····················· | ···················· | ···················· | ···················· |
| N | 3797 | hCoV-19/USA/MN-QDX-2123/2020|EPI_ISL_604480|2020-0 | ···················· | ···················· | ····················· | ···················· | ···················· | ···················· |
| N | 3798 | hCoV-19/USA/MO-QDX-2079/2020|EPI_ISL_604481|2020-0 | ···················· | ···················· | ····················· | ···················· | ···················· | ···················· |
| N | 3799 | hCoV-19/USA/NV-QDX-2082/2020|EPI_ISL_604482|2020-0 | ···················· | ···················· | ····················· | ···················· | ···················· | ···················· |
| N | 3800 | hCoV-19/USA/NV-QDX-2104/2020|EPI_ISL_604483|2020-0 | ···················· | ···················· | ····················· | ···················· | ···················· | ···················· |
| N | 3801 | hCoV-19/USA/NY-QDX-2125/2020|EPI_ISL_604484|2020-0 | ···················· | ···················· | ····················· | ···················· | ···················· | ···················· |
| N | 3802 | hCoV-19/USA/TX-QDX-2111/2020|EPI_ISL_604485|2020-0 | ···················· | ···················· | ····················· | ···················· | ···················· | ···················· |
| N | 3803 | hCoV-19/USA/TX-QDX-2122/2020|EPI_ISL_604486|2020-0 | ···················· | ···················· | ····················· | ···················· | ···················· | ···················· |
| N | 3804 | hCoV-19/USA/TX-QDX-2128/2020|EPI_ISL_604487|2020-0 | ···················· | ···················· | ····················· | ···················· | ···················· | ···················· |
| N | 3805 | hCoV-19/USA/TX-QDX-2151/2020|EPI_ISL_604488|2020-0 | ···················· | ···················· | ····················· | ···················· | ···················· | ···················· |
| N | 3806 | hCoV-19/USA/UT-QDX-2081/2020|EPI_ISL_604489|2020-0 | ···················· | ···················· | ····················· | ···················· | ···················· | ···················· |
| N | 3807 | hCoV-19/USA/UT-QDX-2090/2020|EPI_ISL_604490|2020-0 | ···················· | ···················· | ····················· | ···················· | ···················· | ···················· |
| N | 3808 | hCoV-19/USA/UT-QDX-2103/2020|EPI_ISL_604491|2020-0 | ···················· | ···················· | ····················· | ···················· | ···················· | ···················· |
| N | 3809 | hCoV-19/USA/UT-QDX-2112/2020|EPI_ISL_604492|2020-0 | ···················· | ···················· | ····················· | ···················· | ···················· | ···················· |
| N | 3810 | hCoV-19/USA/UT-QDX-2117/2020|EPI_ISL_604493|2020-0 | ···················· | ···················· | ····················· | ···················· | ···················· | ···················· |
| N | 3811 | hCoV-19/USA/UT-QDX-2126/2020|EPI_ISL_604494|2020-0 | ···················· | ···················· | ····················· | ···················· | ···················· | ···················· |
| N | 3812 | hCoV-19/USA/UT-QDX-2129/2020|EPI_ISL_604495|2020-0 | ···················· | ···················· | ····················· | ···················· | ···················· | ···················· |
| N | 3813 | hCoV-19/USA/UT-QDX-2139/2020|EPI_ISL_604496|2020-0 | ···················· | ···················· | ····················· | ···················· | ···················· | ···················· |
| N | 3814 | hCoV-19/USA/UT-QDX-2167/2020|EPI_ISL_604497|2020-0 | ···················· | ···················· | ····················· | ···················· | ···················· | ···················· |
| N | 3815 | hCoV-19/USA/UT-QDX-2173/2020|EPI_ISL_604498|2020-0 | ···················· | ···················· | ····················· | ···················· | ···················· | ···················· |
| N | 3816 | hCoV-19/USA/WA-QDX-2176/2020|EPI_ISL_604499|2020-0 | ···················· | ···················· | ····················· | ···················· | ···················· | ···················· |
| N | 3817 | hCoV-19/USA/WI-UW-1901/2020|EPI_ISL_605770|2020-09 | ···················· | ···················· | ····················· | ···················· | ···················· | ···················· |
| N | 3818 | hCoV-19/USA/WI-UW-1902/2020|EPI_ISL_605771|2020-09 | ···················· | ···················· | ····················· | ···················· | ···················· | ···················· |
| N | 3819 | hCoV-19/USA/IL-UW-1903/2020|EPI_ISL_605772|2020-09 | ···················· | ···················· | ····················· | ···················· | ···················· | ···················· |
| N | 3820 | hCoV-19/USA/WI-UW-1904/2020|EPI_ISL_605773|2020-09 | ···················· | ···················· | ····················· | ···················· | ···················· | ···················· |
| N | 3821 | hCoV-19/USA/WI-UW-1905/2020|EPI_ISL_605774|2020-09 | ···················· | ···················· | ····················· | ···················· | ···················· | ···················· |
| N | 3823 | hCoV-19/USA/WI-UW-1908/2020|EPI_ISL_605777|2020-09 | ···················· | ···················· | ····················· | ···················· | ···················· | ···················· |
| N | 3824 | hCoV-19/USA/WI-UW-1909/2020|EPI_ISL_605778|2020-09 | ···················· | ···················· | ····················· | ···················· | ···················· | ···················· |
| N | 3825 | hCoV-19/USA/WI-UW-1910/2020|EPI_ISL_605779|2020-09 | ···················· | ···················· | ····················· | ···················· | ···················· | ···················· |
| N | 3826 | hCoV-19/Egypt/C-CEIRS-19 MOH/2020|EPI_ISL_605780|2 | ···················· | ···················· | ····················· | ···················· | ···················· | ···················· |
| N | 3827 | hCoV-19/Bangladesh/JUST-GC91.124/2020|EPI_ISL_6057 | ···················· | ···················· | ····················· | ···················· | ···················· | ···················· |
| N | 3828 | hCoV-19/South Africa/KRISP-K003678/2020|EPI_ISL_60 | ···················· | ···················· | ····················· | ···················· | ···················· | ···················· |
| N | 3829 | hCoV-19/South Africa/KRISP-K003682/2020|EPI_ISL_60 | ···················· | ···················· | ····················· | ···················· | ···················· | ···················· |
| N | 3830 | hCoV-19/South Africa/KRISP-K003720/2020|EPI_ISL_60 | ···················· | ···················· | ····················· | ···················· | ···················· | ···················· |
| N | 3831 | hCoV-19/South Africa/KRISP-K003721/2020|EPI_ISL_60 | ···················· | ···················· | ····················· | ···················· | ···················· | ···················· |
| N | 3832 | hCoV-19/South Africa/KRISP-K003744/2020|EPI_ISL_60 | ···················· | ···················· | ····················· | ···················· | ···················· | ···················· |
| N | 3833 | hCoV-19/USA/DC-BBI2/2020|EPI_ISL_605791|2020-10-14 | ···················· | ···················· | ····················· | ···················· | ···················· | ···················· |
| N | 3834 | hCoV-19/Russia/NVS-RII-34320S/2020|EPI_ISL_605792| | ···················· | ···················· | ····················· | ···················· | ···················· | ···················· |
| N | 3835 | hCoV-19/Russia/Nsk-FRCFTM-1-15a/2020|EPI_ISL_60579 | ···················· | ···················· | ····················· | ···················· | ···················· | ···················· |
| N | 3836 | hCoV-19/Russia/Nsk-FRCFTM-1-16a/2020|EPI_ISL_60579 | ···················· | ···················· | ····················· | ···················· | ···················· | ···················· |
| N | 3837 | hCoV-19/Russia/Nsk-FRCFTM-1-17a/2020|EPI_ISL_60579 | ···················· | ···················· | ····················· | ···················· | ···················· | ···················· |
| N | 3838 | hCoV-19/Russia/Nsk-FRCFTM-1-18a/2020|EPI_ISL_60579 | ···················· | ···················· | ····················· | ···················· | ···················· | ···················· |
| N | 3839 | hCoV-19/Ireland/KY-NVRL-20G35628/2020|EPI_ISL_6057 | ···················· | ···················· | ····················· | ···················· | ···················· | ···················· |
| N | 3840 | hCoV-19/Ireland/D-NVRL-20G35446/2020|EPI_ISL_60579 | ···················· | ·············A······ | ····················· | ···················· | ···················· | ···················· |
| N | 3841 | hCoV-19/England/ALDP-A51640/2020|EPI_ISL_598101|20 | ···················· | ···················· | ····················· | ···················· | ···················· | ···················· |
| N | 3842 | hCoV-19/England/ALDP-A51C02/2020|EPI_ISL_598102|20 | ···················· | ···················· | ····················· | ···················· | ···················· | ···················· |
| N | 3843 | hCoV-19/England/ALDP-A5208D/2020|EPI_ISL_598103|20 | ···················· | ···················· | ····················· | ···················· | ···················· | ···················· |
| N | 3844 | hCoV-19/England/ALDP-A520D8/2020|EPI_ISL_598104|20 | ···················· | ···················· | ····················· | ···················· | ···················· | ···················· |
| N | 3845 | hCoV-19/England/ALDP-A52612/2020|EPI_ISL_598105|20 | ···················· | ·A·······A·········· | ····················· | ···················· | ···················· | ···················· |
| N | 3846 | hCoV-19/England/ALDP-A51570/2020|EPI_ISL_598106|20 | ···················· | ···················· | ····················· | ···················· | ···················· | ···················· |
| N | 3847 | hCoV-19/England/ALDP-A5273D/2020|EPI_ISL_598107|20 | ···················· | ···················· | ····················· | ···················· | ···················· | ···················· |
| N | 3848 | hCoV-19/England/ALDP-A5131C/2020|EPI_ISL_598108|20 | ···················· | ···················· | ····················· | ···················· | ···················· | ···················· |
| N | 3849 | hCoV-19/England/ALDP-A512F1/2020|EPI_ISL_598109|20 | ···················· | ·A·················· | ····················· | ···················· | ···················· | ···················· |
| N | 3850 | hCoV-19/England/ALDP-A51552/2020|EPI_ISL_598110|20 | ···················· | ···················· | ····················· | ···················· | ···················· | ···················· |
| N | 3851 | hCoV-19/England/ALDP-A511A9/2020|EPI_ISL_598111|20 | ···················· | ···················· | ····················· | ···················· | ···················· | ···················· |
| N | 3852 | hCoV-19/England/ALDP-A52366/2020|EPI_ISL_598112|20 | ···················· | ···················· | ····················· | ···················· | ···················· | ···················· |
| N | 3853 | hCoV-19/England/ALDP-A5107F/2020|EPI_ISL_598113|20 | ···················· | ···················· | ····················· | ···················· | ···················· | ···················· |
| N | 3854 | hCoV-19/England/ALDP-A52867/2020|EPI_ISL_598114|20 | ···················· | ···················· | ····················· | ···················· | ···················· | ···················· |
| N | 3855 | hCoV-19/England/ALDP-A514BF/2020|EPI_ISL_598115|20 | ···················· | ·A·················· | ····················· | ···················· | ···················· | ···················· |
| N | 3856 | hCoV-19/England/ALDP-A52348/2020|EPI_ISL_598116|20 | ···················· | ···················· | ····················· | ···················· | ···················· | ···················· |
| N | 3857 | hCoV-19/England/ALDP-A52409/2020|EPI_ISL_598117|20 | ···················· | ···················· | ····················· | ···················· | ···················· | ···················· |

| | | | | | | | | |
|---|---|---|---|---|---|---|---|---|
| N | 3858 | hCoV-19/England/ALDP-A52050/2020\|EPI_ISL_598118\|20 | ····················· | ····················· | ······················ | ····················· | ······T·············· | ····················· |
| N | 3859 | hCoV-19/England/ALDP-A51DC3/2020\|EPI_ISL_598119\|20 | ····················· | ····················· | ······················ | ····················· | ····················· | ····················· |
| N | 3860 | hCoV-19/England/ALDP-A51B23/2020\|EPI_ISL_598120\|20 | ····················· | ····················· | ······················ | ····················· | ····················· | ····················· |
| N | 3861 | hCoV-19/England/ALDP-A516AA/2020\|EPI_ISL_598121\|20 | ····················· | ····················· | ······················ | ····················· | ····················· | ····················· |
| N | 3862 | hCoV-19/England/ALDP-A51B05/2020\|EPI_ISL_598122\|20 | ····················· | ····················· | ······················ | ····················· | ····················· | ····················· |
| N | 3863 | hCoV-19/England/ALDP-A51D69/2020\|EPI_ISL_598123\|20 | ····················· | ····················· | ······················ | ····················· | ····················· | ····················· |
| N | 3864 | hCoV-19/England/ALDP-A51E75/2020\|EPI_ISL_598124\|20 | ····················· | ····················· | ······················ | ····················· | ····················· | ····················· |
| N | 3865 | hCoV-19/England/ALDP-A51A26/2020\|EPI_ISL_598125\|20 | ····················· | ····················· | ······················ | ····················· | ····················· | ····················· |
| N | 3866 | hCoV-19/England/ALDP-A51D4B/2020\|EPI_ISL_598126\|20 | ····················· | ····················· | ······················ | ····················· | ····················· | ····················· |
| N | 3867 | hCoV-19/England/QEUH-A0B2AB/2020\|EPI_ISL_599400\|20 | ····················· | ····················· | ······················ | ····················· | ····················· | ····················· |
| N | 3868 | hCoV-19/England/QEUH-A0BFC2/2020\|EPI_ISL_599401\|20 | ····················· | ····················· | ······················ | ····················· | ····················· | ····················· |
| N | 3869 | hCoV-19/Scotland/QEUH-A0B593/2020\|EPI_ISL_599403\|2 | ····················· | ····················· | ······················ | ····················· | ····················· | ····················· |
| N | 3870 | hCoV-19/Scotland/QEUH-A0BC34/2020\|EPI_ISL_599404\|2 | ····················· | ····················· | ······················ | ····················· | ····················· | ····················· |
| N | 3871 | hCoV-19/England/QEUH-A0B997/2020\|EPI_ISL_599405\|20 | ····················· | ····················· | ······················ | ····················· | ····················· | ····················· |
| N | 3872 | hCoV-19/England/QEUH-A0BA94/2020\|EPI_ISL_599406\|20 | ····················· | ····················· | ······················ | ····················· | ····················· | ····················· |
| N | 3873 | hCoV-19/England/QEUH-A0AC71/2020\|EPI_ISL_599407\|20 | ····················· | ····················· | ······················ | ····················· | ····················· | ····················· |
| N | 3874 | hCoV-19/Scotland/QEUH-A0AE02/2020\|EPI_ISL_599408\|2 | ····················· | ····················· | ······················ | ····················· | ······T·············· | ···············A····· |
| N | 3875 | hCoV-19/Scotland/QEUH-A0BC43/2020\|EPI_ISL_599409\|2 | ····················· | ····················· | ······················ | ····················· | ····················· | ····················· |
| N | 3876 | hCoV-19/England/QEUH-A0BF86/2020\|EPI_ISL_599410\|20 | ····················· | ····················· | ······················ | ····················· | ····················· | ····················· |
| N | 3877 | hCoV-19/England/QEUH-A0C8B7/2020\|EPI_ISL_599411\|20 | ····················· | ····················· | ······················ | ····················· | ····················· | ····················· |
| N | 3878 | hCoV-19/Scotland/QEUH-A0C301/2020\|EPI_ISL_599414\|2 | ····················· | ····················· | ······················ | ····················· | ····················· | ····················· |
| N | 3879 | hCoV-19/Scotland/QEUH-A0B760/2020\|EPI_ISL_599415\|2 | ····················· | ····················· | ······················ | ····················· | ····················· | ····················· |
| N | 3880 | hCoV-19/Scotland/QEUH-A0C37A/2020\|EPI_ISL_599416\|2 | ····················· | ····················· | ······················ | ····················· | ····················· | ····················· |
| N | 3881 | hCoV-19/England/QEUH-A0B3C6/2020\|EPI_ISL_599417\|20 | ····················· | ····················· | ······················ | ····················· | ····················· | ····················· |
| N | 3882 | hCoV-19/England/QEUH-A0AC80/2020\|EPI_ISL_599418\|20 | ····················· | ·A··················· | ······················ | ····················· | ····················· | ····················· |
| N | 3883 | hCoV-19/England/QEUH-A0AD7E/2020\|EPI_ISL_599419\|20 | ····················· | ····················· | ······················ | ····················· | ····················· | ····················· |
| N | 3884 | hCoV-19/England/QEUH-A0B690/2020\|EPI_ISL_599420\|20 | ····················· | ····················· | ······················ | ····················· | ····················· | ····················· |
| N | 3885 | hCoV-19/England/QEUH-A0C046/2020\|EPI_ISL_599421\|20 | ····················· | ····················· | ······················ | ····················· | ····················· | ····················· |
| N | 3886 | hCoV-19/England/QEUH-A0AF78/2020\|EPI_ISL_599423\|20 | ····················· | ····················· | ······················ | ····················· | ····················· | ····················· |
| N | 3887 | hCoV-19/England/QEUH-A0C055/2020\|EPI_ISL_599424\|20 | ····················· | ····················· | ······················ | ····················· | ····················· | ····················· |
| N | 3888 | hCoV-19/England/QEUH-A0B00B/2020\|EPI_ISL_599425\|20 | ····················· | ····················· | ······················ | ····················· | ····················· | ····················· |
| N | 3889 | hCoV-19/Scotland/QEUH-A0B557/2020\|EPI_ISL_599426\|2 | ····················· | ····················· | ······················ | ····················· | ····················· | ····················· |
| N | 3890 | hCoV-19/England/QEUH-A0C064/2020\|EPI_ISL_599427\|20 | ····················· | ····················· | ······················ | ····················· | ····················· | ····················· |
| N | 3891 | hCoV-19/England/QEUH-A0BAEF/2020\|EPI_ISL_599428\|20 | ····················· | ····················· | ······················ | ····················· | ····················· | ····················· |
| N | 3892 | hCoV-19/Scotland/QEUH-A0BCBC/2020\|EPI_ISL_599429\|2 | ····················· | ····················· | ······················ | ····················· | ····················· | ····················· |
| N | 3893 | hCoV-19/England/QEUH-A0BB91/2020\|EPI_ISL_599430\|20 | ····················· | ····················· | ······················ | ····················· | ····················· | ····················· |
| N | 3894 | hCoV-19/Scotland/QEUH-A0C29B/2020\|EPI_ISL_599431\|2 | ····················· | ····················· | ······················ | ····················· | ····················· | ····················· |
| N | 3895 | hCoV-19/England/QEUH-A0AAC2/2020\|EPI_ISL_599432\|20 | ····················· | ····················· | ······················ | ····················· | ····················· | ····················· |
| N | 3896 | hCoV-19/England/QEUH-A0AFA5/2020\|EPI_ISL_599433\|20 | ····················· | ····················· | ······················ | ····················· | ····················· | ····················· |
| N | 3897 | hCoV-19/England/QEUH-A0C8A8/2020\|EPI_ISL_599435\|20 | ····················· | ····················· | ······················ | ····················· | ····················· | ····················· |
| N | 3898 | hCoV-19/England/QEUH-A0C8C6/2020\|EPI_ISL_599436\|20 | ····················· | ····················· | ······················ | ····················· | ····················· | ····················· |
| N | 3899 | hCoV-19/Scotland/QEUH-A0B821/2020\|EPI_ISL_599437\|2 | ····················· | ····················· | ······················ | ····················· | ····················· | ····················· |
| N | 3900 | hCoV-19/England/QEUH-A0BA0D/2020\|EPI_ISL_599438\|20 | ····················· | ····················· | ······················ | ····················· | ····················· | ····················· |
| N | 3901 | hCoV-19/England/QEUH-A0BE6B/2020\|EPI_ISL_599439\|20 | ····················· | ····················· | ······················ | ····················· | ····················· | ····················· |
| N | 3902 | hCoV-19/England/QEUH-A0C86C/2020\|EPI_ISL_599440\|20 | ····················· | ····················· | ······················ | ····················· | ····················· | ····················· |
| N | 3903 | hCoV-19/England/QEUH-A0AEF3/2020\|EPI_ISL_599441\|20 | ····················· | ····················· | ······················ | ····················· | ····················· | ····················· |
| N | 3904 | hCoV-19/England/QEUH-A0B074/2020\|EPI_ISL_599442\|20 | ····················· | ····················· | ······················ | ····················· | ····················· | ····················· |
| N | 3905 | hCoV-19/England/QEUH-A0BDB9/2020\|EPI_ISL_599443\|20 | ····················· | ·A··················· | ······················ | ····················· | ····················· | ···A················· |
| N | 3906 | hCoV-19/England/QEUH-A0AD6F/2020\|EPI_ISL_599444\|20 | ····················· | ····················· | ······················ | ····················· | ····················· | ····················· |
| N | 3907 | hCoV-19/Scotland/QEUH-A0C36B/2020\|EPI_ISL_599445\|2 | ····················· | ····················· | ······················ | ····················· | ····················· | ····················· |
| N | 3908 | hCoV-19/England/QEUH-A0AFB4/2020\|EPI_ISL_599446\|20 | ····················· | ····················· | ······················ | ····················· | ····················· | ····················· |
| N | 3909 | hCoV-19/England/QEUH-A0ACBD/2020\|EPI_ISL_599447\|20 | ····················· | ····················· | ······················ | ····················· | ····················· | ····················· |
| N | 3910 | hCoV-19/England/QEUH-A0B126/2020\|EPI_ISL_599448\|20 | ····················· | ····················· | ······················ | ····················· | ····················· | ····················· |
| N | 3911 | hCoV-19/Scotland/QEUH-A0B79D/2020\|EPI_ISL_599449\|2 | ····················· | ····················· | ······················ | ····················· | ····················· | ····················· |
| N | 3912 | hCoV-19/Scotland/QEUH-A0BCAD/2020\|EPI_ISL_599450\|2 | ····················· | ····················· | ······················ | ····················· | ····················· | ····················· |
| N | 3913 | hCoV-19/England/QEUH-A0AED5/2020\|EPI_ISL_599451\|20 | ····················· | ····················· | ······················ | ····················· | ····················· | ····················· |
| N | 3914 | hCoV-19/Scotland/QEUH-A0BEC5/2020\|EPI_ISL_599452\|2 | ····················· | ····················· | ······················ | ····················· | ····················· | ····················· |
| N | 3915 | hCoV-19/England/QEUH-A0BBDD/2020\|EPI_ISL_599453\|20 | ····················· | ····················· | ······················ | ····················· | ····················· | ····················· |
| N | 3916 | hCoV-19/England/QEUH-A0AAA4/2020\|EPI_ISL_599454\|20 | ····················· | ····················· | ······················ | ····················· | ····················· | ····················· |
| N | 3917 | hCoV-19/England/QEUH-A0C398/2020\|EPI_ISL_599455\|20 | ····················· | ····················· | ······················ | ····················· | ····················· | ····················· |
| N | 3918 | hCoV-19/England/QEUH-A0B162/2020\|EPI_ISL_599456\|20 | ····················· | ····················· | ······················ | ····················· | ····················· | ····················· |
| N | 3919 | hCoV-19/Scotland/QEUH-A0ADE7/2020\|EPI_ISL_599457\|2 | ····················· | ····················· | ······················ | ····················· | ····················· | ····················· |
| N | 3920 | hCoV-19/Scotland/QEUH-9F392A/2020\|EPI_ISL_600800\|2 | ····················· | ····················· | ······················ | ····················· | ····················· | ····················· |
| N | 3921 | hCoV-19/England/QEUH-9F3061/2020\|EPI_ISL_600801\|20 | ····················· | ·A··················· | ······················ | ····················· | ····················· | ····················· |
| N | 3922 | hCoV-19/Scotland/QEUH-9F3641/2020\|EPI_ISL_600802\|2 | ····················· | ····················· | ······················ | ····················· | ····················· | ····················· |
| N | 3923 | hCoV-19/England/QEUH-9F3CD6/2020\|EPI_ISL_600803\|20 | ····················· | ····················· | ······················ | ····················· | ····················· | ····················· |
| N | 3924 | hCoV-19/Scotland/QEUH-9F30CB/2020\|EPI_ISL_600804\|2 | ····················· | ····················· | ······················ | ····················· | ····················· | ····················· |
| N | 3925 | hCoV-19/Scotland/QEUH-9F2D2F/2020\|EPI_ISL_600805\|2 | ····················· | ····················· | ······················ | ····················· | ····················· | ····················· |
| N | 3926 | hCoV-19/England/QEUH-9F308F/2020\|EPI_ISL_600806\|20 | ····················· | ····················· | ······················ | ····················· | ······T·············· | ····················· |
| N | 3927 | hCoV-19/Scotland/QEUH-9F3F82/2020\|EPI_ISL_600807\|2 | ····················· | ····················· | ······················ | ····················· | ····················· | ····················· |
| N | 3928 | hCoV-19/England/QEUH-9F3FA0/2020\|EPI_ISL_600810\|20 | ····················· | ····················· | ······················ | ····················· | ····················· | ····················· |
| N | 3929 | hCoV-19/Scotland/QEUH-9F3140/2020\|EPI_ISL_600811\|2 | ····················· | ····················· | ······················ | ····················· | ····················· | ····················· |
| N | 3930 | hCoV-19/Scotland/QEUH-9F2CD7/2020\|EPI_ISL_600812\|2 | ····················· | ····················· | ······················ | ····················· | ····················· | ····················· |
| N | 3931 | hCoV-19/Scotland/QEUH-9F3BF7/2020\|EPI_ISL_600813\|2 | ····················· | ·A··················· | ······················ | ····················· | ····················· | ····················· |
| N | 3932 | hCoV-19/England/QEUH-9F2C04/2020\|EPI_ISL_600814\|20 | ····················· | ····················· | ······················ | ····················· | ····················· | ····················· |
| N | 3933 | hCoV-19/Scotland/QEUH-9F3A90/2020\|EPI_ISL_600815\|2 | ····················· | ····················· | ······················ | ····················· | ····················· | ····················· |
| N | 3934 | hCoV-19/Scotland/QEUH-9F3C12/2020\|EPI_ISL_600816\|2 | ····················· | ····················· | ······················ | ····················· | ····················· | ····················· |
| N | 3935 | hCoV-19/Scotland/QEUH-9F2E95/2020\|EPI_ISL_600817\|2 | ····················· | ····················· | ······················ | ····················· | ····················· | ····················· |
| N | 3936 | hCoV-19/Scotland/QEUH-9F3D3D/2020\|EPI_ISL_600819\|2 | ····················· | ····················· | ······················ | ····················· | ····················· | ····················· |
| N | 3937 | hCoV-19/Scotland/QEUH-9F41D6/2020\|EPI_ISL_600820\|2 | ····················· | ····················· | ······················ | ····················· | ····················· | ····················· |

| | | | | | | | |
|---|---|---|---|---|---|---|---|
| N | 3938 hCoV-19/Scotland/QEUH-9F3D79/2020\|EPI_ISL_600821\|2 | ···················· | ···················· | ···················· | ···················· | ···················· | ···················· |
| N | 3939 hCoV-19/England/QEUH-9F3070/2020\|EPI_ISL_600822\|20 | ···················· | ···················· | ···················· | ···················· | ·····T·············· | ···················· |
| N | 3940 hCoV-19/Scotland/QEUH-9F3D2E/2020\|EPI_ISL_600823\|2 | ···················· | ···················· | ···················· | ···················· | ···················· | ···················· |
| N | 3941 hCoV-19/Scotland/QEUH-9F4297/2020\|EPI_ISL_600825\|2 | ···················· | ···················· | ···················· | ···················· | ···················· | ···················· |
| N | 3942 hCoV-19/England/QEUH-9F423D/2020\|EPI_ISL_600826\|20 | ···················· | ···················· | ···················· | ···················· | ···················· | ···················· |
| N | 3943 hCoV-19/Scotland/QEUH-9F30DA/2020\|EPI_ISL_600827\|2 | ···················· | ···················· | ···················· | ···················· | ···················· | ···················· |
| N | 3944 hCoV-19/England/QEUH-9F2BF8/2020\|EPI_ISL_600828\|20 | ···················· | ···················· | ···················· | ···················· | ···················· | ···················· |
| N | 3945 hCoV-19/Scotland/QEUH-9F84AC/2020\|EPI_ISL_600829\|2 | ···················· | ···················· | ···················· | ···················· | ···················· | ···················· |
| N | 3946 hCoV-19/Scotland/QEUH-9F59DD/2020\|EPI_ISL_600830\|2 | ···················· | ···················· | ···················· | ···················· | ···················· | ···················· |
| N | 3947 hCoV-19/Scotland/QEUH-9F8503/2020\|EPI_ISL_600831\|2 | ···················· | ···················· | ···················· | ···················· | ···················· | ···················· |
| N | 3948 hCoV-19/Scotland/QEUH-9F5F71/2020\|EPI_ISL_600832\|2 | ···················· | ···················· | ···················· | ···················· | ···················· | ···················· |
| N | 3949 hCoV-19/Scotland/QEUH-9F848E/2020\|EPI_ISL_600833\|2 | ···················· | ···················· | ···················· | ···················· | ···················· | ···················· |
| N | 3950 hCoV-19/Scotland/QEUH-9F620E/2020\|EPI_ISL_600834\|2 | ···················· | ···················· | ···················· | ···················· | ···················· | ···················· |
| N | 3951 hCoV-19/Scotland/QEUH-9F61D4/2020\|EPI_ISL_600835\|2 | ···················· | ···················· | ···················· | ···················· | ···················· | ···················· |
| N | 3952 hCoV-19/Scotland/QEUH-9F6875/2020\|EPI_ISL_600836\|2 | ···················· | ···················· | ···················· | ···················· | ···················· | ···················· |
| N | 3953 hCoV-19/Scotland/QEUH-9F612F/2020\|EPI_ISL_600837\|2 | ···················· | ···················· | ···················· | ···················· | ···················· | ···················· |
| N | 3954 hCoV-19/Scotland/QEUH-9F5788/2020\|EPI_ISL_600838\|2 | ···················· | ···················· | ···················· | ···················· | ···················· | ···················· |
| N | 3955 hCoV-19/Scotland/QEUH-9F858B/2020\|EPI_ISL_600839\|2 | ···················· | ···················· | ···················· | ···················· | ···················· | ···················· |
| N | 3956 hCoV-19/Scotland/QEUH-9F84CA/2020\|EPI_ISL_600840\|2 | ···················· | ···················· | ···················· | ···················· | ···················· | ···················· |
| N | 3957 hCoV-19/Scotland/QEUH-9F84BB/2020\|EPI_ISL_600841\|2 | ···················· | ···················· | ···················· | ···················· | ···················· | ···················· |
| N | 3958 hCoV-19/Scotland/QEUH-9F8DDE/2020\|EPI_ISL_600842\|2 | ···················· | ···················· | ···················· | ···················· | ···················· | ···················· |
| N | 3959 hCoV-19/Scotland/QEUH-9F90B6/2020\|EPI_ISL_600843\|2 | ···················· | ···················· | ···················· | ···················· | ···················· | ···················· |
| N | 3960 hCoV-19/Scotland/QEUH-9F8521/2020\|EPI_ISL_600844\|2 | ···················· | ···················· | ···················· | ···················· | ···················· | ···················· |
| N | 3961 hCoV-19/Scotland/QEUH-9F8DFC/2020\|EPI_ISL_600845\|2 | ···················· | ···················· | ···················· | ···················· | ···················· | ···················· |
| N | 3962 hCoV-19/Scotland/QEUH-9F67B4/2020\|EPI_ISL_600846\|2 | ···················· | ···················· | ···················· | ···················· | ···················· | ···················· |
| N | 3963 hCoV-19/Scotland/QEUH-9F5621/2020\|EPI_ISL_600847\|2 | ···················· | ···················· | ···················· | ···················· | ···················· | ···················· |
| N | 3964 hCoV-19/Scotland/QEUH-9F854F/2020\|EPI_ISL_600848\|2 | ···················· | ···················· | ···················· | ···················· | ···················· | ···················· |
| N | 3965 hCoV-19/Scotland/QEUH-9F61F2/2020\|EPI_ISL_600849\|2 | ···················· | ···················· | ···················· | ···················· | ···················· | ···················· |
| N | 3966 hCoV-19/Scotland/QEUH-9F614D/2020\|EPI_ISL_600850\|2 | ···················· | ···················· | ···················· | ···················· | ···················· | ···················· |
| N | 3967 hCoV-19/Scotland/QEUH-9F847F/2020\|EPI_ISL_600852\|2 | ···················· | ···················· | ···················· | ···················· | ···················· | ···················· |
| N | 3968 hCoV-19/Scotland/QEUH-9F6198/2020\|EPI_ISL_600853\|2 | ···················· | ···················· | ···················· | ···················· | ···················· | ···················· |
| N | 3969 hCoV-19/Scotland/QEUH-9F682A/2020\|EPI_ISL_600854\|2 | ···················· | ···················· | ···················· | ···················· | ···················· | ···················· |
| N | 3970 hCoV-19/Wales/QEUH-9F5FDB/2020\|EPI_ISL_600855\|2020 | ···················· | ···················· | ···················· | ···················· | ···················· | ···················· |
| N | 3971 hCoV-19/Scotland/QEUH-9F572E/2020\|EPI_ISL_600856\|2 | ···················· | ···················· | ···················· | ···················· | ···················· | ···················· |
| N | 3972 hCoV-19/Scotland/QEUH-9F615C/2020\|EPI_ISL_600857\|2 | ···················· | ···················· | ···················· | ···················· | ···················· | ···················· |
| N | 3973 hCoV-19/Scotland/QEUH-9F680C/2020\|EPI_ISL_600858\|2 | ···················· | ···················· | ···················· | ···················· | ···················· | ···················· |
| N | 3974 hCoV-19/Scotland/QEUH-9F855E/2020\|EPI_ISL_600859\|2 | ···················· | ···················· | ···················· | ···················· | ···················· | ···················· |
| N | 3975 hCoV-19/Scotland/QEUH-9F84E8/2020\|EPI_ISL_600860\|2 | ···················· | ···················· | ···················· | ···················· | ···················· | ···················· |
| N | 3976 hCoV-19/Scotland/QEUH-9F59FB/2020\|EPI_ISL_600861\|2 | ···················· | ···················· | ···················· | ···················· | ···················· | ···················· |
| N | 3977 hCoV-19/Scotland/QEUH-9F59EC/2020\|EPI_ISL_600862\|2 | ···················· | ···················· | ···················· | ···················· | ···················· | ···················· |
| N | 3978 hCoV-19/Scotland/QEUH-9F859A/2020\|EPI_ISL_600863\|2 | ···················· | ···················· | ···················· | ···················· | ···················· | ···················· |
| N | 3979 hCoV-19/Scotland/QEUH-9F84F7/2020\|EPI_ISL_600865\|2 | ···················· | ···················· | ···················· | ···················· | ···················· | ···················· |
| N | 3980 hCoV-19/Scotland/QEUH-9F8DED/2020\|EPI_ISL_600866\|2 | ···················· | ···················· | ···················· | ···················· | ···················· | ···················· |
| N | 3981 hCoV-19/Scotland/QEUH-9F85A9/2020\|EPI_ISL_600867\|2 | ···················· | ···················· | ···················· | ···················· | ···················· | ···················· |
| N | 3982 hCoV-19/Scotland/QEUH-9F565E/2020\|EPI_ISL_600868\|2 | ···················· | ···················· | ···················· | ···················· | ···················· | ···················· |
| N | 3983 hCoV-19/Scotland/QEUH-9F6189/2020\|EPI_ISL_600869\|2 | ···················· | ···················· | ···················· | ···················· | ···················· | ···················· |
| N | 3984 hCoV-19/Scotland/QEUH-9F61B6/2020\|EPI_ISL_600870\|2 | ···················· | ···················· | ···················· | ···················· | ·····T·············· | ···················· |
| N | 3985 hCoV-19/Scotland/QEUH-9F564F/2020\|EPI_ISL_600871\|2 | ···················· | ···················· | ···················· | ···················· | ···················· | ···················· |
| N | 3986 hCoV-19/Scotland/QEUH-9F6893/2020\|EPI_ISL_600872\|2 | ···················· | ···················· | ···················· | ···················· | ···················· | ···················· |
| N | 3987 hCoV-19/Scotland/QEUH-9F6110/2020\|EPI_ISL_600873\|2 | ···················· | ···················· | ···················· | ···················· | ···················· | ···················· |
| N | 3988 hCoV-19/Scotland/QEUH-9F8E26/2020\|EPI_ISL_600874\|2 | ···················· | ···················· | ···················· | ···················· | ···················· | ···················· |
| N | 3989 hCoV-19/Scotland/QEUH-9F61C5/2020\|EPI_ISL_600875\|2 | ···················· | ···················· | ···················· | ···················· | ···················· | ···················· |
| N | 3990 hCoV-19/Scotland/QEUH-9F61E3/2020\|EPI_ISL_600876\|2 | ···················· | ···················· | ···················· | ···················· | ···················· | ···················· |
| N | 3991 hCoV-19/Scotland/QEUH-9F856D/2020\|EPI_ISL_600878\|2 | ···················· | ···················· | ···················· | ···················· | ···················· | ···················· |
| N | 3992 hCoV-19/Scotland/QEUH-9F8530/2020\|EPI_ISL_600879\|2 | ···················· | ···················· | ···················· | ···················· | ···················· | ···················· |
| N | 3993 hCoV-19/Scotland/QEUH-9F67A5/2020\|EPI_ISL_600880\|2 | ···················· | ···················· | ···················· | ···················· | ···················· | ···················· |
| N | 3994 hCoV-19/Scotland/QEUH-9F8DCF/2020\|EPI_ISL_600881\|2 | ···················· | ·A·················· | ···················· | ···················· | ···················· | ···················· |
| N | 3995 hCoV-19/Scotland/QEUH-9F8E44/2020\|EPI_ISL_600883\|2 | ···················· | ···················· | ···················· | ···················· | ···················· | ···················· |
| N | 3996 hCoV-19/Scotland/QEUH-9F616B/2020\|EPI_ISL_600884\|2 | ···················· | ···················· | ···················· | ···················· | ···················· | ···················· |
| N | 3997 hCoV-19/Scotland/QEUH-9F9405/2020\|EPI_ISL_600886\|2 | ···················· | ···················· | ···················· | ···················· | ···················· | ···················· |
| N | 3998 hCoV-19/Scotland/QEUH-9F8451/2020\|EPI_ISL_600887\|2 | ···················· | ···················· | ···················· | ···················· | ···················· | ···················· |
| N | 3999 hCoV-19/Scotland/QEUH-9F617A/2020\|EPI_ISL_600888\|2 | ···················· | ···················· | ···················· | ···················· | ···················· | ···················· |
| N | 4000 hCoV-19/Scotland/QEUH-9F5FAE/2020\|EPI_ISL_600889\|2 | ···················· | ···················· | ···················· | ···················· | ···················· | ···················· |
| N | 4001 hCoV-19/Scotland/QEUH-9F7416/2020\|EPI_ISL_600890\|2 | ···················· | ···················· | ···················· | ···················· | ···················· | ···················· |
| N | 4002 hCoV-19/Scotland/QEUH-9F94AB/2020\|EPI_ISL_600891\|2 | ···················· | ···················· | ···················· | ···················· | ···················· | ···················· |
| N | 4003 hCoV-19/Wales/MILK-9F2387/2020\|EPI_ISL_600892\|2020 | ···················· | ···················· | ···················· | ···················· | ···················· | ···················· |
| N | 4004 hCoV-19/Scotland/QEUH-9F9951/2020\|EPI_ISL_600893\|2 | ···················· | ···················· | ···················· | ···················· | ···················· | ···················· |
| N | 4005 hCoV-19/Scotland/QEUH-9F94C9/2020\|EPI_ISL_600894\|2 | ···················· | ···················· | ···················· | ···················· | ···················· | ···················· |
| N | 4006 hCoV-19/Scotland/QEUH-9F7DA2/2020\|EPI_ISL_600895\|2 | ···················· | ···················· | ···················· | ···················· | ···················· | ···················· |
| N | 4007 hCoV-19/Scotland/QEUH-9F7D57/2020\|EPI_ISL_600896\|2 | ···················· | ···················· | ···················· | ···················· | ···················· | ···················· |
| N | 4008 hCoV-19/Wales/QEUH-9F9678/2020\|EPI_ISL_600897\|2020 | ···················· | ···················· | ···················· | ···················· | ···················· | ···················· |
| N | 4009 hCoV-19/Scotland/QEUH-9F7D2A/2020\|EPI_ISL_600898\|2 | ···················· | ···················· | ···················· | ···················· | ···················· | ···················· |
| N | 4010 hCoV-19/Scotland/QEUH-9F94D8/2020\|EPI_ISL_600899\|2 | ···················· | ···················· | ···················· | ···················· | ···················· | ···················· |
| N | 4011 hCoV-19/England/QEUH-9DBFCC/2020\|EPI_ISL_601250\|20 | ···················· | ···················· | ···················· | ···················· | ···················· | ···················· |
| N | 4012 hCoV-19/England/QEUH-9DA552/2020\|EPI_ISL_601251\|20 | ···················· | ···················· | ···················· | ···················· | ···················· | ···················· |
| N | 4013 hCoV-19/England/QEUH-9DA8A4/2020\|EPI_ISL_601252\|20 | ···················· | ···················· | ···················· | ···················· | ···················· | ···················· |
| N | 4014 hCoV-19/England/QEUH-9DA69B/2020\|EPI_ISL_601253\|20 | ···················· | ···················· | ···················· | ···················· | ···················· | ···················· |
| N | 4015 hCoV-19/England/QEUH-9DA534/2020\|EPI_ISL_601254\|20 | ···················· | ···················· | ···················· | ···················· | ···················· | ···················· |
| N | 4016 hCoV-19/England/MILK-9E03F5/2020\|EPI_ISL_601256\|20 | ···················· | ···················· | ···················· | ···················· | ···················· | ···················· |
| N | 4017 hCoV-19/England/QEUH-9DB041/2020\|EPI_ISL_601257\|20 | ···················· | ···················· | ···················· | ···················· | ···················· | ···················· |

| | | | | | | | | |
|---|---|---|---|---|---|---|---|---|
| N | 4018 | hCoV-19/England/MILK-9E0586/2020\|EPI_ISL_601258\|20 | ···················· | ·A·················· | ···················· | ···················· | ···················· | ···················· |
| N | 4019 | hCoV-19/England/QEUH-9DA91A/2020\|EPI_ISL_601259\|20 | ···················· | ···················· | ···················· | ···················· | ···················· | ···················· |
| N | 4020 | hCoV-19/England/QEUH-9DA604/2020\|EPI_ISL_601260\|20 | ···················· | ···················· | ···················· | ···················· | ···················· | ···················· |
| N | 4021 | hCoV-19/England/QEUH-9DBEED/2020\|EPI_ISL_601261\|20 | ···················· | ···················· | ···················· | ···················· | ···················· | ···················· |
| N | 4022 | hCoV-19/Wales/MILK-9E0261/2020\|EPI_ISL_601263\|2020 | ···················· | ···················· | ···················· | ···················· | ···················· | ···················· |
| N | 4023 | hCoV-19/Wales/MILK-9E04F2/2020\|EPI_ISL_601264\|2020 | ···················· | ···················· | ···················· | ···················· | ···················· | ···················· |
| N | 4024 | hCoV-19/England/QEUH-9DC2D1/2020\|EPI_ISL_601265\|20 | ···················· | ···················· | ···················· | ···················· | ···················· | ···················· |
| N | 4025 | hCoV-19/England/QEUH-9DBD4A/2020\|EPI_ISL_601266\|20 | ···················· | ·A·················· | ···················· | ···················· | ···················· | ··A················· |
| N | 4026 | hCoV-19/England/MILK-9E086F/2020\|EPI_ISL_601267\|20 | ···················· | ···················· | ···················· | ···················· | ···················· | ···················· |
| N | 4027 | hCoV-19/England/MILK-9E029E/2020\|EPI_ISL_601268\|20 | ···················· | ···················· | ···················· | ···················· | ···················· | ···················· |
| N | 4028 | hCoV-19/England/QEUH-9DA6C8/2020\|EPI_ISL_601269\|20 | ···················· | ·A·················· | ···················· | ···················· | ···················· | ··A················· |
| N | 4029 | hCoV-19/England/MILK-9E042F/2020\|EPI_ISL_601270\|20 | ···················· | ···················· | ···················· | ···················· | ···················· | ···················· |
| N | 4030 | hCoV-19/England/QEUH-9DC13E/2020\|EPI_ISL_601271\|20 | ···················· | ···················· | ···················· | ···················· | ···················· | ···················· |
| N | 4031 | hCoV-19/England/QEUH-9DBE56/2020\|EPI_ISL_601274\|20 | ···················· | ···················· | ···················· | ···················· | ···················· | ···················· |
| N | 4032 | hCoV-19/England/MILK-9E0313/2020\|EPI_ISL_601275\|20 | ···················· | ···················· | ···················· | ···················· | ···················· | ···················· |
| N | 4033 | hCoV-19/England/QEUH-9DBE74/2020\|EPI_ISL_601276\|20 | ···················· | ···················· | ···················· | ···················· | ···················· | ···················· |
| N | 4034 | hCoV-19/Wales/MILK-9E093F/2020\|EPI_ISL_601277\|2020 | ···················· | ···················· | ···················· | ···················· | ···················· | ···················· |
| N | 4035 | hCoV-19/England/QEUH-9DC30B/2020\|EPI_ISL_601279\|20 | ···················· | ···················· | ···················· | ···················· | ···················· | ···················· |
| N | 4036 | hCoV-19/Australia/SAP499/2020\|EPI_ISL_602580\|2020- | ···················· | ·A·················· | ···················· | ···················· | ···················· | ···················· |
| N | 4037 | hCoV-19/Australia/SAP500/2020\|EPI_ISL_602581\|2020- | ···················· | ·A·················· | ···················· | ···················· | ···················· | ···················· |
| N | 4038 | hCoV-19/Lithuania/AL-LUHS-Eilnr1/2020\|EPI_ISL_6030 | ···················· | ···················· | ···················· | ···················· | ···················· | ···················· |
| N | 4039 | hCoV-19/Lithuania/AL-LUHS-Eilnr2/2020\|EPI_ISL_6030 | ···················· | ···················· | ···················· | ···················· | ···················· | ···················· |
| N | 4040 | hCoV-19/Lithuania/KU-LUHS-Eilnr4/2020\|EPI_ISL_6030 | ···················· | ···················· | ···················· | ···················· | ···················· | ···················· |
| N | 4041 | hCoV-19/Lithuania/KU-LUHS-Eilnr10/2020\|EPI_ISL_603 | ···················· | ···················· | ···················· | ···················· | ···················· | ···················· |
| N | 4042 | hCoV-19/Lithuania/MR-LUHS-Eilnr15/2020\|EPI_ISL_603 | ···················· | ···················· | ···················· | ···················· | ···················· | ···················· |
| N | 4043 | hCoV-19/Lithuania/MR-LUHS-Eilnr19/2020\|EPI_ISL_603 | ···················· | ···················· | ···················· | ···················· | ···················· | ···················· |
| N | 4044 | hCoV-19/Lithuania/KU-LUHS-Eilnr22/2020\|EPI_ISL_603 | ···················· | ···················· | ···················· | ···················· | ···················· | ···················· |
| N | 4045 | hCoV-19/Lithuania/KU-LUHS-Eilnr24/2020\|EPI_ISL_603 | ···················· | ···················· | ···················· | ···················· | ···················· | ···················· |
| N | 4046 | hCoV-19/Lithuania/MR-LUHS-Eilnr30/2020\|EPI_ISL_603 | ···················· | ···················· | ···················· | ···················· | ···················· | ···················· |
| N | 4047 | hCoV-19/Lithuania/MR-LUHS-Eilnr32/2020\|EPI_ISL_603 | ···················· | ···················· | ···················· | ···················· | ···················· | ···················· |
| N | 4048 | hCoV-19/Lithuania/MR-LUHS-Eilnr38/2020\|EPI_ISL_603 | ···················· | ···················· | ···················· | ···················· | ···················· | ···················· |
| N | 4049 | hCoV-19/Lithuania/MR-LUHS-Eilnr39/2020\|EPI_ISL_603 | ···················· | ···················· | ···················· | ···················· | ···················· | ···················· |
| N | 4050 | hCoV-19/Lithuania/MR-LUHS-Eilnr47/2020\|EPI_ISL_603 | ···················· | ···················· | ···················· | ···················· | ···················· | ···················· |
| N | 4051 | hCoV-19/Lithuania/KL-LUHS-Eilnr59/2020\|EPI_ISL_603 | ···················· | ···················· | ···················· | ···················· | ···················· | ···················· |
| N | 4052 | hCoV-19/Lithuania/KU-LUHS-Eilnr61/2020\|EPI_ISL_603 | ···················· | ···················· | ···················· | ···················· | ···················· | ···················· |
| N | 4053 | hCoV-19/Lithuania/MR-LUHS-Eilnr67/2020\|EPI_ISL_603 | ···················· | ···················· | ···················· | ···················· | ···················· | ···················· |
| N | 4054 | hCoV-19/Lithuania/MR-LUHS-Eilnr69/2020\|EPI_ISL_603 | ···················· | ···················· | ···················· | ···················· | ···················· | ···················· |
| N | 4055 | hCoV-19/Lithuania/MR-LUHS-Eilnr71/2020\|EPI_ISL_603 | ···················· | ···················· | ···················· | ···················· | ···················· | ···················· |
| N | 4056 | hCoV-19/Italy/LAZ-INMI-33/2020\|EPI_ISL_603140\|2020 | ···················· | ···················· | ···················· | ···················· | ···················· | ···················· |
| N | 4057 | hCoV-19/Italy/LAZ-INMI-34/2020\|EPI_ISL_603141\|2020 | ···················· | ···················· | ···················· | ···················· | ···················· | ···················· |
| N | 4058 | hCoV-19/Italy/LAZ-INMI-35/2020\|EPI_ISL_603142\|2020 | ···················· | ···················· | ···················· | ···················· | ···················· | ···················· |
| N | 4059 | hCoV-19/Italy/LAZ-INMI-36/2020\|EPI_ISL_603143\|2020 | ···················· | ···················· | ···················· | ···················· | ···················· | ···················· |
| N | 4060 | hCoV-19/Italy/LAZ-INMI-37/2020\|EPI_ISL_603144\|2020 | ···················· | ···················· | ···················· | ···················· | ···················· | ···················· |
| N | 4061 | hCoV-19/Italy/LAZ-INMI-38/2020\|EPI_ISL_603145\|2020 | ···················· | ···················· | ···················· | ···················· | ···················· | ···················· |
| N | 4062 | hCoV-19/Italy/LAZ-INMI-41/2020\|EPI_ISL_603148\|2020 | ···················· | ···················· | ···················· | ···················· | ···················· | ···················· |
| N | 4063 | hCoV-19/Italy/LAZ-INMI-42/2020\|EPI_ISL_603149\|2020 | ···················· | ···················· | ···················· | ···················· | ···················· | ···················· |
| N | 4064 | hCoV-19/USA/MI-QDX-2161/2020\|EPI_ISL_604470\|2020-0 | ···················· | ···················· | ···················· | ···················· | ···················· | ···················· |
| N | 4065 | hCoV-19/USA/MI-QDX-2164/2020\|EPI_ISL_604471\|2020-0 | ···················· | ···················· | ···················· | ···················· | ···················· | ···················· |
| N | 4066 | hCoV-19/USA/MI-QDX-2168/2020\|EPI_ISL_604472\|2020-0 | ···················· | ···················· | ···················· | ···················· | ···················· | ···················· |
| N | 4067 | hCoV-19/USA/MI-QDX-2172/2020\|EPI_ISL_604473\|2020-0 | ···················· | ···················· | ···················· | ···················· | ···················· | ···················· |
| N | 4068 | hCoV-19/USA/MI-QDX-2178/2020\|EPI_ISL_604474\|2020-0 | ···················· | ···················· | ···················· | ···················· | ···················· | ···················· |
| N | 4069 | hCoV-19/USA/MI-QDX-2182/2020\|EPI_ISL_604475\|2020-0 | ···················· | ···················· | ···················· | ···················· | ···················· | ···················· |
| N | 4070 | hCoV-19/USA/MI-QDX-2183/2020\|EPI_ISL_604476\|2020-0 | ···················· | ···················· | ···················· | ···················· | ···················· | ···················· |
| N | 4071 | hCoV-19/USA/MI-QDX-2186/2020\|EPI_ISL_604478\|2020-0 | ···················· | ···················· | ···················· | ···················· | ···················· | ···················· |
| N | 4072 | hCoV-19/USA/MI-QDX-2187/2020\|EPI_ISL_604479\|2020-0 | ···················· | ···················· | ···················· | ···················· | ···················· | ···················· |
| N | 4073 | hCoV-19/Hong Kong/HKPU-5135/2020\|EPI_ISL_610201\|20 | ···················· | ·A·················· | ···················· | ···················· | ···················· | ··A················· |
| N | 4074 | hCoV-19/Hong Kong/HKPU-5138/2020\|EPI_ISL_610202\|20 | ···················· | ·A·················· | ···················· | ···················· | ···················· | ··A················· |
| N | 4075 | hCoV-19/Hong Kong/HKPU-5136/2020\|EPI_ISL_610203\|20 | ···················· | ·A·················· | ···················· | ···················· | ···················· | ··A················· |
| N | 4076 | hCoV-19/Hong Kong/HKPU-5111/2020\|EPI_ISL_610204\|20 | ···················· | ·A·················· | ···················· | ···················· | ···················· | ··A················· |
| N | 4077 | hCoV-19/Hong Kong/HKPU-4789/2020\|EPI_ISL_610205\|20 | ···················· | ···················· | ···················· | ···················· | ···················· | ···················· |
| N | 4078 | hCoV-19/Hong Kong/HKPU-4278/2020\|EPI_ISL_610206\|20 | ···················· | ···················· | ···················· | ···················· | ···················· | ···················· |
| N | 4079 | hCoV-19/Hong Kong/HKPU-4328/2020\|EPI_ISL_610207\|20 | ···················· | ···················· | ···················· | ···················· | ···················· | ···················· |
| N | 4080 | hCoV-19/Hong Kong/HKPU-3138/2020\|EPI_ISL_610208\|20 | ···················· | ···················· | ···················· | ···················· | ···················· | ···················· |
| N | 4081 | hCoV-19/Hong Kong/HKPU-2514/2020\|EPI_ISL_610209\|20 | ···················· | ···················· | ···················· | ···················· | ···················· | ···················· |
| N | 4082 | hCoV-19/Hong Kong/HKPU-1142/2020\|EPI_ISL_610210\|20 | ···················· | ···················· | ···················· | ···················· | ···················· | ···················· |
| N | 4084 | hCoV-19/Hong Kong/HKPU-3034/2020\|EPI_ISL_610212\|20 | ···················· | ···················· | ···················· | ···················· | ···················· | ···················· |
| N | 4085 | hCoV-19/Hong Kong/HKPU-4424/2020\|EPI_ISL_610213\|20 | ···················· | ···················· | ···················· | ···················· | ···················· | ···················· |
| N | 4086 | hCoV-19/Hong Kong/HKPU-5126/2020\|EPI_ISL_610214\|20 | ···················· | ·A·················· | ···················· | ···················· | ···················· | ··A················· |
| N | 4087 | hCoV-19/Hong Kong/HKPU-5102/2020\|EPI_ISL_610215\|20 | ···················· | ·A·················· | ···················· | ···················· | ···················· | ··A················· |
| N | 4090 | hCoV-19/Hong Kong/HKPU-4973/2020\|EPI_ISL_610219\|20 | ···················· | ·A·················· | ···················· | ···················· | ···················· | ···················· |
| N | 4091 | hCoV-19/Hong Kong/HKPU-4999/2020\|EPI_ISL_610220\|20 | ···················· | ···················· | ···················· | ···················· | ···················· | ···················· |
| N | 4093 | hCoV-19/Hong Kong/HKPU-5132/2020\|EPI_ISL_610222\|20 | ···················· | ·A·················· | ···················· | ···················· | ···················· | ··A················· |
| N | 4096 | hCoV-19/Hong Kong/HKPU-5178/2020\|EPI_ISL_610225\|20 | ···················· | ···················· | ···················· | ···················· | ···················· | ···················· |
| N | 4097 | hCoV-19/England/QEUH-A5E7C8/2020\|EPI_ISL_597500\|20 | ···················· | ···················· | ···················· | ···················· | ···················· | ···················· |
| N | 4098 | hCoV-19/England/QEUH-A5E151/2020\|EPI_ISL_597501\|20 | ···················· | ···················· | ···················· | ···················· | ···················· | ···················· |
| N | 4099 | hCoV-19/England/CAMC-A57659/2020\|EPI_ISL_597502\|20 | ···················· | ···················· | ···················· | ···················· | ···················· | ···················· |
| N | 4100 | hCoV-19/England/QEUH-A5E5FB/2020\|EPI_ISL_597503\|20 | ···················· | ···················· | ···················· | ···················· | ···················· | ···················· |
| N | 4101 | hCoV-19/England/CAMC-A56F8C/2020\|EPI_ISL_597504\|20 | ···················· | ···················· | ···················· | ···················· | ···················· | ···················· |
| N | 4102 | hCoV-19/England/CAMC-A573AD/2020\|EPI_ISL_597505\|20 | ···················· | ···················· | ···················· | ···················· | ······T············· | ···················· |
| N | 4103 | hCoV-19/England/QEUH-A5E397/2020\|EPI_ISL_597506\|20 | ···················· | ···················· | ···················· | ···················· | ···················· | ···················· |

| | | | | | | | | |
|---|---|---|---|---|---|---|---|---|
| N | 4104 | hCoV-19/England/CAMC-A57783/2020\|EPI_ISL_597507\|20 | ···················· | ···················· | ····················· | ···················· | ···················· | ···················· |
| N | 4105 | hCoV-19/England/CAMC-A571B2/2020\|EPI_ISL_597508\|20 | ···················· | ···················· | ····················· | ···················· | ···················· | ···················· |
| N | 4106 | hCoV-19/England/QEUH-A5E379/2020\|EPI_ISL_597509\|20 | ···················· | ···················· | ····················· | ···················· | ···················· | ···················· |
| N | 4107 | hCoV-19/England/CAMC-A57808/2020\|EPI_ISL_597510\|20 | ···················· | ···················· | ····················· | ···················· | ···················· | ···················· |
| N | 4108 | hCoV-19/England/ALDP-A4FE3E/2020\|EPI_ISL_597555\|20 | ···················· | ···················· | ····················· | ···················· | ···················· | ···················· |
| N | 4109 | hCoV-19/England/ALDP-A4FD04/2020\|EPI_ISL_597556\|20 | ···················· | ···················· | ····················· | ···················· | ···················· | ···················· |
| N | 4110 | hCoV-19/England/ALDP-A50AEB/2020\|EPI_ISL_597557\|20 | ···················· | ·A·················· | ····················· | ···················· | ···················· | ···················· |
| N | 4111 | hCoV-19/England/ALDP-A5033B/2020\|EPI_ISL_597558\|20 | ···················· | ···················· | ····················· | ···················· | ···················· | ···················· |
| N | 4112 | hCoV-19/England/ALDP-A50034/2020\|EPI_ISL_597559\|20 | ···················· | ·A·················· | ····················· | ···················· | ···················· | ···A················ |
| N | 4113 | hCoV-19/England/ALDP-A5009E/2020\|EPI_ISL_597560\|20 | ···················· | ···················· | ····················· | ···················· | ···················· | ···················· |
| N | 4114 | hCoV-19/England/ALDP-A50BCA/2020\|EPI_ISL_597561\|20 | ···················· | ···················· | ····················· | ···················· | ···················· | ···················· |
| N | 4115 | hCoV-19/England/ALDP-A4FCCB/2020\|EPI_ISL_597562\|20 | ···················· | ···················· | ····················· | ···················· | ···················· | ···················· |
| N | 4116 | hCoV-19/England/ALDP-A5024D/2020\|EPI_ISL_597563\|20 | ···················· | ···················· | ····················· | ···················· | ···················· | ···················· |
| N | 4117 | hCoV-19/England/ALDP-A50298/2020\|EPI_ISL_597565\|20 | ···················· | ···················· | ····················· | ···················· | ···················· | ···················· |
| N | 4118 | hCoV-19/England/ALDP-A50A72/2020\|EPI_ISL_597566\|20 | ···················· | ·A·················· | ····················· | ···················· | ···················· | ···A················ |
| N | 4119 | hCoV-19/England/ALDP-A505F9/2020\|EPI_ISL_597567\|20 | ···················· | ···················· | ····················· | ···················· | ···················· | ···················· |
| N | 4120 | hCoV-19/England/ALDP-A501C8/2020\|EPI_ISL_597568\|20 | ···················· | ···················· | ····················· | ···················· | ···················· | ···················· |
| N | 4121 | hCoV-19/England/ALDP-A50D88/2020\|EPI_ISL_597569\|20 | ···················· | ···················· | ····················· | ···················· | ···················· | ···················· |
| N | 4122 | hCoV-19/England/ALDP-A50025/2020\|EPI_ISL_597570\|20 | ···················· | ·A·················· | ····················· | ···················· | ···················· | ···················· |
| N | 4123 | hCoV-19/England/ALDP-A4FC9E/2020\|EPI_ISL_597572\|20 | ···················· | ···················· | ····················· | ···················· | ···················· | ···················· |
| N | 4124 | hCoV-19/England/ALDP-A4FBEC/2020\|EPI_ISL_597573\|20 | ···················· | ···················· | ····················· | ···················· | ···················· | ···················· |
| N | 4125 | hCoV-19/England/ALDP-A4FB28/2020\|EPI_ISL_597574\|20 | ···················· | ···················· | ····················· | ···················· | ···················· | ···················· |
| N | 4126 | hCoV-19/England/ALDP-A50ABE/2020\|EPI_ISL_597576\|20 | ···················· | ···················· | ····················· | ···················· | ···················· | ···················· |
| N | 4127 | hCoV-19/England/ALDP-A510E8/2020\|EPI_ISL_597577\|20 | ···················· | ···················· | ····················· | ···················· | ···················· | ···················· |
| N | 4128 | hCoV-19/England/ALDP-A50122/2020\|EPI_ISL_597578\|20 | ···················· | ···················· | ····················· | ···················· | ···················· | ···················· |
| N | 4129 | hCoV-19/England/ALDP-A4FA0D/2020\|EPI_ISL_597579\|20 | ···················· | ···················· | ····················· | ···················· | ···················· | ···················· |
| N | 4130 | hCoV-19/England/ALDP-A50447/2020\|EPI_ISL_597580\|20 | ···················· | ···················· | ····················· | ···················· | ···················· | ···················· |
| N | 4131 | hCoV-19/England/ALDP-A500E9/2020\|EPI_ISL_597581\|20 | ···················· | ···················· | ····················· | ···················· | ···················· | ···················· |
| N | 4132 | hCoV-19/England/ALDP-A50DE2/2020\|EPI_ISL_597582\|20 | ···················· | ···················· | ····················· | ···················· | ···················· | ···················· |
| N | 4133 | hCoV-19/England/ALDP-A4FD8C/2020\|EPI_ISL_597583\|20 | ···················· | ···················· | ····················· | ···················· | ···················· | ···················· |
| N | 4134 | hCoV-19/England/ALDP-A50456/2020\|EPI_ISL_597584\|20 | ···················· | ···················· | ····················· | ···················· | ···················· | ···················· |
| N | 4135 | hCoV-19/England/ALDP-A50D97/2020\|EPI_ISL_597585\|20 | ···················· | ···················· | ····················· | ···················· | ···················· | ···················· |
| N | 4136 | hCoV-19/England/ALDP-A4FFD1/2020\|EPI_ISL_597586\|20 | ···················· | ···················· | ····················· | ···················· | ···················· | ···················· |
| N | 4137 | hCoV-19/England/ALDP-A507C6/2020\|EPI_ISL_597587\|20 | ···················· | ···················· | ····················· | ···················· | ···················· | ···················· |
| N | 4138 | hCoV-19/England/ALDP-A50878/2020\|EPI_ISL_597588\|20 | ···················· | ···················· | ····················· | ···················· | ···················· | ···················· |
| N | 4139 | hCoV-19/England/ALDP-A4FC34/2020\|EPI_ISL_597589\|20 | ···················· | ···················· | ····················· | ···················· | ···················· | ···················· |
| N | 4140 | hCoV-19/England/ALDP-A4FAA3/2020\|EPI_ISL_597590\|20 | ···················· | ···················· | ····················· | ···················· | ···················· | ···················· |
| N | 4141 | hCoV-19/England/ALDP-A4FC07/2020\|EPI_ISL_597591\|20 | ···················· | ···················· | ····················· | ···················· | ···················· | ···················· |
| N | 4142 | hCoV-19/England/ALDP-A4FB91/2020\|EPI_ISL_597592\|20 | ···················· | ···················· | ····················· | ···················· | ···················· | ···················· |
| N | 4143 | hCoV-19/England/ALDP-A4FF4A/2020\|EPI_ISL_597593\|20 | ···················· | ···················· | ····················· | ···················· | ···················· | ···················· |
| N | 4144 | hCoV-19/England/ALDP-A4FA94/2020\|EPI_ISL_597594\|20 | ···················· | ···················· | ····················· | ···················· | ···················· | ···················· |
| N | 4145 | hCoV-19/England/ALDP-A50D79/2020\|EPI_ISL_597595\|20 | ···················· | ···················· | ····················· | ···················· | ···················· | ···················· |
| N | 4146 | hCoV-19/England/ALDP-A50113/2020\|EPI_ISL_597596\|20 | ···················· | ···················· | ····················· | ···················· | ···················· | ···················· |
| N | 4147 | hCoV-19/England/ALDP-A50623/2020\|EPI_ISL_597597\|20 | ···················· | ···················· | ····················· | ···················· | ···················· | ···················· |
| N | 4148 | hCoV-19/England/ALDP-A50C9A/2020\|EPI_ISL_597598\|20 | ···················· | ·A·················· | ····················· | ···················· | ···················· | ···A················ |
| N | 4149 | hCoV-19/England/ALDP-A5076C/2020\|EPI_ISL_597599\|20 | ···················· | ···················· | ····················· | ···················· | ···················· | ···················· |
| N | 4150 | hCoV-19/England/MILK-A44B14/2020\|EPI_ISL_598800\|20 | ···················· | ···················· | ····················· | ···················· | ···················· | ···················· |
| N | 4151 | hCoV-19/England/MILK-A44BAB/2020\|EPI_ISL_598801\|20 | ···················· | ···················· | ····················· | ···················· | ···················· | ···················· |
| N | 4152 | hCoV-19/England/MILK-A45EFC/2020\|EPI_ISL_598802\|20 | ···················· | ···················· | ····················· | ···················· | ···················· | ···················· |
| N | 4153 | hCoV-19/England/MILK-A45F08/2020\|EPI_ISL_598803\|20 | ···················· | ···················· | ····················· | ···················· | ···················· | ···················· |
| N | 4154 | hCoV-19/England/MILK-A45FAE/2020\|EPI_ISL_598805\|20 | ···················· | ···················· | ····················· | ···················· | ······T············· | ···················· |
| N | 4155 | hCoV-19/England/MILK-A468B1/2020\|EPI_ISL_598806\|20 | ···················· | ···················· | ····················· | ···················· | ···················· | ···················· |
| N | 4156 | hCoV-19/England/MILK-A459BF/2020\|EPI_ISL_598807\|20 | ···················· | ···················· | ····················· | ···················· | ···················· | ···················· |
| N | 4157 | hCoV-19/England/MILK-A456E5/2020\|EPI_ISL_598809\|20 | ···················· | ···················· | ····················· | ···················· | ···················· | ···················· |
| N | 4158 | hCoV-19/England/MILK-A44DC3/2020\|EPI_ISL_598818\|20 | ···················· | ···················· | ····················· | ···················· | ···················· | ···················· |
| N | 4159 | hCoV-19/England/MILK-A44EC0/2020\|EPI_ISL_598819\|20 | ···················· | ·A·················· | ····················· | ···················· | ···················· | ···················· |
| N | 4160 | hCoV-19/England/MILK-A44F54/2020\|EPI_ISL_598820\|20 | ···················· | ···················· | ····················· | ···················· | ···················· | ···················· |
| N | 4161 | hCoV-19/England/MILK-A45BC8/2020\|EPI_ISL_598822\|20 | ···················· | ···················· | ····················· | ···················· | ···················· | ···················· |
| N | 4162 | hCoV-19/England/MILK-A44EA2/2020\|EPI_ISL_598823\|20 | ···················· | ···················· | ····················· | ···················· | ······T············· | ···················· |
| N | 4163 | hCoV-19/Wales/ALDP-A2D49E/2020\|EPI_ISL_598824\|2020 | ···················· | ···················· | ····················· | ···················· | ···················· | ···················· |
| N | 4164 | hCoV-19/Wales/ALDP-A2D57D/2020\|EPI_ISL_598825\|2020 | ···················· | ···················· | ····················· | ···················· | ···················· | ···················· |
| N | 4165 | hCoV-19/Wales/ALDP-A2D540/2020\|EPI_ISL_598826\|2020 | ···················· | ···················· | ····················· | ···················· | ···················· | ···················· |
| N | 4166 | hCoV-19/Wales/ALDP-A2EF23/2020\|EPI_ISL_598827\|2020 | ···················· | ···················· | ····················· | ···················· | ···················· | ···················· |
| N | 4167 | hCoV-19/England/ALDP-A2F098/2020\|EPI_ISL_598828\|20 | ···················· | ···················· | ····················· | ···················· | ···················· | ···················· |
| N | 4168 | hCoV-19/Wales/ALDP-A2EEDB/2020\|EPI_ISL_598829\|2020 | ···················· | ···················· | ····················· | ···················· | ···················· | ···················· |
| N | 4169 | hCoV-19/Wales/ALDP-A2FECB/2020\|EPI_ISL_598830\|2020 | ···················· | ···················· | ····················· | ···················· | ···················· | ···················· |
| N | 4170 | hCoV-19/Scotland/QEUH-A211C0/2020\|EPI_ISL_598831\|2 | ···················· | ···················· | ····················· | ···················· | ······T············· | ···················· |
| N | 4171 | hCoV-19/Scotland/QEUH-A1EA52/2020\|EPI_ISL_598832\|2 | ···················· | ···················· | ····················· | ···················· | ···················· | ···················· |
| N | 4172 | hCoV-19/Scotland/QEUH-A2195F/2020\|EPI_ISL_598833\|2 | ···················· | ···················· | ····················· | ···················· | ···················· | ···················· |
| N | 4173 | hCoV-19/Scotland/QEUH-A21F6C/2020\|EPI_ISL_598834\|2 | ···················· | ···················· | ····················· | ···················· | ···················· | ···················· |
| N | 4174 | hCoV-19/Scotland/QEUH-A1D9FC/2020\|EPI_ISL_598835\|2 | ···················· | ···················· | ····················· | ···················· | ···················· | ···················· |
| N | 4175 | hCoV-19/Scotland/QEUH-A1E3DF/2020\|EPI_ISL_598836\|2 | ···················· | ···················· | ····················· | ···················· | ···················· | ···················· |
| N | 4176 | hCoV-19/Scotland/QEUH-A21658/2020\|EPI_ISL_598837\|2 | ···················· | ···················· | ····················· | ···················· | ···················· | ···················· |
| N | 4177 | hCoV-19/England/QEUH-A1E937/2020\|EPI_ISL_598838\|20 | ···················· | ···················· | ····················· | ···················· | ···················· | ···················· |
| N | 4178 | hCoV-19/Scotland/QEUH-A217DD/2020\|EPI_ISL_598839\|2 | ···················· | ···················· | ····················· | ···················· | ···················· | ···················· |
| N | 4179 | hCoV-19/Scotland/QEUH-A1E3EE/2020\|EPI_ISL_598840\|2 | ···················· | ···················· | ····················· | ···················· | ···················· | ···················· |
| N | 4180 | hCoV-19/Scotland/QEUH-A1E445/2020\|EPI_ISL_598841\|2 | ···················· | ···················· | ····················· | ···················· | ···················· | ···················· |
| N | 4181 | hCoV-19/Scotland/QEUH-A217FB/2020\|EPI_ISL_598886\|2 | ···················· | ···················· | ····················· | ···················· | ···················· | ···················· |
| N | 4182 | hCoV-19/Scotland/QEUH-A21588/2020\|EPI_ISL_598887\|2 | ···················· | ···················· | ····················· | ···················· | ···················· | ···················· |
| N | 4183 | hCoV-19/Scotland/QEUH-A1E0AB/2020\|EPI_ISL_598888\|2 | ···················· | ···················· | ····················· | ···················· | ···················· | ···················· |

| | | | | | | | | |
|---|---|---|---|---|---|---|---|---|
| N | 4184 | hCoV-19/Scotland/QEUH-A218E9/2020\|EPI_ISL_598889\|2 | .................... | .................... | .................... | .................... | .................... | .................... |
| N | 4185 | hCoV-19/Scotland/QEUH-A2163A/2020\|EPI_ISL_598891\|2 | .................... | .................... | .................... | .................... | .................... | .................... |
| N | 4186 | hCoV-19/Scotland/QEUH-A1E30C/2020\|EPI_ISL_598892\|2 | .................... | .................... | .................... | .................... | .................... | .................... |
| N | 4187 | hCoV-19/Scotland/QEUH-A2196E/2020\|EPI_ISL_598893\|2 | .................... | .................... | .................... | .................... | .................... | .................... |
| N | 4188 | hCoV-19/Scotland/QEUH-A1EA25/2020\|EPI_ISL_598894\|2 | .................... | .................... | .................... | .................... | .................... | .................... |
| N | 4189 | hCoV-19/Scotland/QEUH-A1E515/2020\|EPI_ISL_598895\|2 | .................... | .................... | .................... | .................... | ......T............. | .................... |
| N | 4190 | hCoV-19/Scotland/QEUH-A1EE56/2020\|EPI_ISL_598896\|2 | .................... | .................... | .................... | .................... | .................... | .................... |
| N | 4191 | hCoV-19/Scotland/QEUH-A1EA70/2020\|EPI_ISL_598897\|2 | .................... | .................... | .................... | .................... | .................... | .................... |
| N | 4192 | hCoV-19/Scotland/QEUH-A1E0E7/2020\|EPI_ISL_598898\|2 | .................... | .................... | .................... | .................... | .................... | .................... |
| N | 4193 | hCoV-19/Scotland/QEUH-A1E07E/2020\|EPI_ISL_598899\|2 | .................... | .................... | .................... | .................... | .................... | .................... |
| N | 4194 | hCoV-19/Scotland/QEUH-9DED47/2020\|EPI_ISL_601200\|2 | .................... | .................... | .................... | .................... | .................... | .................... |
| N | 4195 | hCoV-19/Scotland/QEUH-9DE6F1/2020\|EPI_ISL_601201\|2 | .................... | .................... | .................... | .................... | .................... | .................... |
| N | 4196 | hCoV-19/Scotland/QEUH-9DEDB0/2020\|EPI_ISL_601202\|2 | .................... | .................... | .................... | .................... | .................... | .................... |
| N | 4197 | hCoV-19/Scotland/QEUH-9DE697/2020\|EPI_ISL_601204\|2 | .................... | .................... | .................... | .................... | .................... | .................... |
| N | 4198 | hCoV-19/Scotland/QEUH-9DEAE6/2020\|EPI_ISL_601205\|2 | .................... | .................... | .................... | .................... | .................... | .................... |
| N | 4199 | hCoV-19/Scotland/QEUH-9DED56/2020\|EPI_ISL_601206\|2 | .................... | .................... | .................... | .................... | .................... | .................... |
| N | 4200 | hCoV-19/Scotland/QEUH-9DEA40/2020\|EPI_ISL_601207\|2 | .................... | .................... | .................... | .................... | .................... | .................... |
| N | 4201 | hCoV-19/Scotland/QEUH-9DEA6E/2020\|EPI_ISL_601208\|2 | .................... | .................... | .................... | .................... | .................... | .................... |
| N | 4202 | hCoV-19/Scotland/QEUH-9DEA04/2020\|EPI_ISL_601209\|2 | .................... | .A.................. | .................... | .................... | .................... | .................... |
| N | 4203 | hCoV-19/Scotland/QEUH-9DEA7D/2020\|EPI_ISL_601210\|2 | .................... | .................... | .................... | .................... | .................... | .................... |
| N | 4204 | hCoV-19/Scotland/QEUH-9DE6D3/2020\|EPI_ISL_601211\|2 | .................... | .................... | .................... | .................... | .................... | .................... |
| N | 4205 | hCoV-19/Scotland/QEUH-9DE9DA/2020\|EPI_ISL_601212\|2 | .................... | .................... | .................... | .................... | .................... | .................... |
| N | 4206 | hCoV-19/Scotland/QEUH-9DEB01/2020\|EPI_ISL_601213\|2 | .................... | .................... | .................... | .................... | .................... | .................... |
| N | 4207 | hCoV-19/Scotland/QEUH-9DEAD7/2020\|EPI_ISL_601215\|2 | .................... | .................... | .................... | .................... | .................... | .................... |
| N | 4208 | hCoV-19/Scotland/QEUH-9DE6E2/2020\|EPI_ISL_601216\|2 | .................... | .................... | .................... | .................... | .................... | .................... |
| N | 4209 | hCoV-19/Scotland/QEUH-9DE6A6/2020\|EPI_ISL_601217\|2 | .................... | .................... | .................... | .................... | .................... | .................... |
| N | 4210 | hCoV-19/Scotland/QEUH-9DEAAA/2020\|EPI_ISL_601218\|2 | .................... | .................... | .................... | .................... | .................... | .................... |
| N | 4211 | hCoV-19/Scotland/QEUH-9DEDA1/2020\|EPI_ISL_601219\|2 | .................... | .................... | .................... | .................... | .................... | .................... |
| N | 4212 | hCoV-19/Scotland/QEUH-9DEA8C/2020\|EPI_ISL_601220\|2 | .................... | .................... | .................... | .................... | .................... | .................... |
| N | 4213 | hCoV-19/Scotland/QEUH-9DE72B/2020\|EPI_ISL_601221\|2 | .................... | .................... | .................... | .................... | .................... | .................... |
| N | 4214 | hCoV-19/Scotland/QEUH-9DEA13/2020\|EPI_ISL_601222\|2 | .................... | .................... | .................... | .................... | .................... | .................... |
| N | 4215 | hCoV-19/Scotland/QEUH-9DE71C/2020\|EPI_ISL_601223\|2 | .................... | .................... | .................... | .................... | .................... | .................... |
| N | 4216 | hCoV-19/Scotland/MILK-9E3116/2020\|EPI_ISL_601224\|2 | .................... | .................... | .................... | .................... | .................... | .................... |
| N | 4217 | hCoV-19/Scotland/QEUH-9DE873/2020\|EPI_ISL_601226\|2 | .................... | .................... | .................... | .................... | .................... | .................... |
| N | 4218 | hCoV-19/Scotland/QEUH-9DEDFC/2020\|EPI_ISL_601227\|2 | .................... | .................... | .................... | .................... | .................... | .................... |
| N | 4219 | hCoV-19/Scotland/QEUH-9DEAF5/2020\|EPI_ISL_601228\|2 | .................... | .................... | .................... | .................... | .................... | .................... |
| N | 4220 | hCoV-19/Scotland/QEUH-9DE7B2/2020\|EPI_ISL_601229\|2 | .................... | .................... | .................... | .................... | .................... | .................... |
| N | 4221 | hCoV-19/Scotland/QEUH-9DEA9B/2020\|EPI_ISL_601230\|2 | .................... | .................... | .................... | .................... | .................... | .................... |
| N | 4222 | hCoV-19/Scotland/QEUH-9DEAB9/2020\|EPI_ISL_601231\|2 | .................... | .................... | .................... | .................... | .................... | .................... |
| N | 4223 | hCoV-19/Scotland/QEUH-9DE8A0/2020\|EPI_ISL_601232\|2 | .................... | .................... | .................... | .................... | .................... | .................... |
| N | 4224 | hCoV-19/Scotland/QEUH-9DEE17/2020\|EPI_ISL_601233\|2 | .................... | .................... | .................... | .................... | .................... | .................... |
| N | 4225 | hCoV-19/England/MILK-9E0B2A/2020\|EPI_ISL_601234\|20 | .................... | .................... | .................... | .................... | .................... | .................... |
| N | 4226 | hCoV-19/England/MILK-9E045C/2020\|EPI_ISL_601235\|20 | .................... | .................... | .................... | .................... | .................... | .................... |
| N | 4227 | hCoV-19/England/QEUH-9DBE65/2020\|EPI_ISL_601236\|20 | .................... | .................... | .................... | .................... | .................... | .................... |
| N | 4228 | hCoV-19/England/QEUH-9DC14D/2020\|EPI_ISL_601237\|20 | .................... | .A.................. | .................... | .................... | .................... | ...A................ |
| N | 4229 | hCoV-19/England/QEUH-9DA974/2020\|EPI_ISL_601238\|20 | .................... | .................... | .................... | .................... | .................... | .................... |
| N | 4230 | hCoV-19/England/MILK-9E0BEE/2020\|EPI_ISL_601239\|20 | .................... | .................... | .................... | .................... | .................... | .................... |
| N | 4231 | hCoV-19/England/MILK-9E0B84/2020\|EPI_ISL_601240\|20 | .................... | .................... | .................... | .................... | .................... | .................... |
| N | 4232 | hCoV-19/England/MILK-9E0CDC/2020\|EPI_ISL_601242\|20 | .................... | .................... | .................... | .................... | .................... | .................... |
| N | 4233 | hCoV-19/Wales/MILK-9E09B7/2020\|EPI_ISL_601243\|2020 | .................... | .................... | .................... | .................... | .................... | .................... |
| N | 4234 | hCoV-19/England/QEUH-9DA4DD/2020\|EPI_ISL_601244\|20 | .................... | .A.................. | .................... | .................... | .................... | .................... |
| N | 4235 | hCoV-19/England/QEUH-9DC24A/2020\|EPI_ISL_601246\|20 | .................... | .................... | .................... | .................... | .................... | .................... |
| N | 4236 | hCoV-19/England/MILK-9E0771/2020\|EPI_ISL_601248\|20 | .................... | .................... | .................... | .................... | .................... | .................... |
| N | 4237 | hCoV-19/England/MILK-9E0C90/2020\|EPI_ISL_601249\|20 | .................... | .................... | .................... | .................... | .................... | .................... |
| N | 4238 | hCoV-19/England/MILK-9E07DB/2020\|EPI_ISL_601272\|20 | .................... | .................... | .................... | .................... | .................... | .................... |
| N | 4239 | hCoV-19/Wales/MILK-9E0902/2020\|EPI_ISL_601273\|2020 | .................... | .................... | .................... | .................... | .................... | .................... |
| N | 4240 | hCoV-19/England/MILK-9E02DA/2020\|EPI_ISL_601280\|20 | .................... | .................... | .................... | .................... | .................... | .................... |
| N | 4241 | hCoV-19/England/MILK-9E079F/2020\|EPI_ISL_601281\|20 | .................... | .................... | .................... | .................... | .................... | .................... |
| N | 4242 | hCoV-19/England/MILK-9E0401/2020\|EPI_ISL_601282\|20 | .................... | .................... | .................... | .................... | .................... | .................... |
| N | 4243 | hCoV-19/Wales/MILK-9E03C8/2020\|EPI_ISL_601283\|2020 | .................... | .................... | .................... | .................... | .................... | .................... |
| N | 4244 | hCoV-19/England/QEUH-9DBDC2/2020\|EPI_ISL_601284\|20 | .................... | .................... | .................... | .................... | .................... | .................... |
| N | 4245 | hCoV-19/England/QEUH-9DA7A7/2020\|EPI_ISL_601285\|20 | .................... | .A.................. | .................... | .................... | .................... | .................... |
| N | 4246 | hCoV-19/England/QEUH-9DA613/2020\|EPI_ISL_601286\|20 | .................... | .................... | .................... | .................... | .................... | .................... |
| N | 4247 | hCoV-19/England/MILK-9E0850/2020\|EPI_ISL_601287\|20 | .................... | .................... | .................... | .................... | .................... | .................... |
| N | 4248 | hCoV-19/England/QEUH-9DC0F5/2020\|EPI_ISL_601288\|20 | .................... | .................... | .................... | .................... | .................... | .................... |
| N | 4249 | hCoV-19/Wales/MILK-9E0D42/2020\|EPI_ISL_601289\|2020 | .................... | .................... | .................... | .................... | .................... | .................... |
| N | 4250 | hCoV-19/England/MILK-9E0B66/2020\|EPI_ISL_601291\|20 | .................... | .A.................. | .................... | .................... | .................... | ...A................ |
| N | 4251 | hCoV-19/Wales/MILK-9E0841/2020\|EPI_ISL_601292\|2020 | .................... | .................... | .................... | .................... | .................... | .................... |
| N | 4252 | hCoV-19/Wales/MILK-9E0216/2020\|EPI_ISL_601293\|2020 | .................... | .................... | .................... | .................... | .................... | .................... |
| N | 4253 | hCoV-19/England/MILK-9E05A4/2020\|EPI_ISL_601294\|20 | .................... | .................... | .................... | .................... | .................... | .................... |
| N | 4254 | hCoV-19/England/QEUH-9DC277/2020\|EPI_ISL_601295\|20 | .................... | .................... | .................... | .................... | .................... | .................... |
| N | 4255 | hCoV-19/England/QEUH-9DC110/2020\|EPI_ISL_601296\|20 | .................... | .................... | .................... | .................... | .................... | .................... |
| N | 4256 | hCoV-19/England/MILK-9E06B0/2020\|EPI_ISL_601298\|20 | .................... | .A.................. | .................... | .................... | .................... | .................... |
| N | 4257 | hCoV-19/England/MILK-9E039B/2020\|EPI_ISL_601299\|20 | .................... | .................... | .................... | .................... | .................... | .................... |
| N | 4258 | hCoV-19/Germany/NW-HHU-178/2020\|EPI_ISL_602500\|202 | .................... | .................... | .................... | .................... | .................... | .................... |
| N | 4259 | hCoV-19/Germany/NW-HHU-179/2020\|EPI_ISL_602501\|202 | .................... | .................... | .................... | .................... | .................... | .................... |
| N | 4260 | hCoV-19/Germany/NW-HHU-180/2020\|EPI_ISL_602502\|202 | .................... | .................... | .................... | .................... | .................... | .................... |
| N | 4261 | hCoV-19/Germany/NW-HHU-181/2020\|EPI_ISL_602503\|202 | .................... | .................... | .................... | .................... | .................... | .................... |
| N | 4262 | hCoV-19/Germany/NW-HHU-182/2020\|EPI_ISL_602504\|202 | .................... | .................... | .................... | .................... | .................... | .................... |
| N | 4263 | hCoV-19/Germany/NW-HHU-183/2020\|EPI_ISL_602505\|202 | .................... | .................... | .................... | .................... | .................... | .................... |

| | | | | | | | | |
|---|---|---|---|---|---|---|---|---|
| N | 4264 | hCoV-19/Germany/NW-HHU-184/2020\|EPI_ISL_602506\|202 | .................... | .................... | .................... | .................... | .................... | .................... |
| N | 4265 | hCoV-19/Germany/NW-HHU-185/2020\|EPI_ISL_602507\|202 | .................... | .................... | .................... | .................... | .................... | .................... |
| N | 4266 | hCoV-19/Germany/NW-HHU-186/2020\|EPI_ISL_602508\|202 | .................... | .................... | .................... | .................... | .................... | .................... |
| N | 4267 | hCoV-19/Germany/NW-HHU-187/2020\|EPI_ISL_602509\|202 | .................... | .................... | .................... | .................... | .................... | .................... |
| N | 4268 | hCoV-19/Germany/NW-HHU-143/2020\|EPI_ISL_602513\|202 | .................... | .................... | .................... | .................... | .................... | .................... |
| N | 4269 | hCoV-19/Germany/NW-HHU-147/2020\|EPI_ISL_602517\|202 | .................... | .................... | .................... | .................... | .................... | .................... |
| N | 4270 | hCoV-19/Germany/NW-HHU-148/2020\|EPI_ISL_602518\|202 | .................... | .................... | .................... | .................... | .................... | .................... |
| N | 4271 | hCoV-19/Germany/NW-HHU-149/2020\|EPI_ISL_602519\|202 | .................... | .................... | .................... | .................... | .................... | .................... |
| N | 4272 | hCoV-19/Germany/NW-HHU-150/2020\|EPI_ISL_602520\|202 | .................... | .................... | .................... | .................... | .................... | .................... |
| N | 4273 | hCoV-19/Germany/NW-HHU-152/2020\|EPI_ISL_602522\|202 | .................... | .................... | .................... | .................... | .................... | .................... |
| N | 4274 | hCoV-19/Germany/NW-HHU-153/2020\|EPI_ISL_602523\|202 | .................... | .................... | .................... | .................... | .................... | .................... |
| N | 4275 | hCoV-19/Germany/NW-HHU-154/2020\|EPI_ISL_602524\|202 | .................... | .................... | .................... | .................... | .................... | .................... |
| N | 4276 | hCoV-19/Germany/NW-HHU-158/2020\|EPI_ISL_602527\|202 | .................... | .................... | .................... | .................... | .................... | .................... |
| N | 4277 | hCoV-19/Germany/NW-HHU-160/2020\|EPI_ISL_602529\|202 | .................... | .................... | .................... | .................... | .................... | .................... |
| N | 4278 | hCoV-19/Germany/NW-HHU-161/2020\|EPI_ISL_602530\|202 | .................... | .................... | .................... | .................... | .................... | .................... |
| N | 4279 | hCoV-19/Sri Lanka/MIN01/2020\|EPI_ISL_602564\|2020-1 | .................... | .................... | .................... | .................... | .................... | .................... |
| N | 4280 | hCoV-19/Australia/SAP493/2020\|EPI_ISL_602577\|2020- | .................... | .................... | .................... | .................... | .................... | .................... |
| N | 4281 | hCoV-19/Australia/SAP498/2020\|EPI_ISL_602579\|2020- | .................... | .................... | .................... | .................... | .................... | .................... |
| N | 4282 | hCoV-19/USA/MA-QDX-1458/2020\|EPI_ISL_603825\|2020-0 | .................... | .................... | .................... | .................... | .................... | .................... |
| N | 4283 | hCoV-19/USA/MA-QDX-1460/2020\|EPI_ISL_603826\|2020-0 | .................... | .................... | .................... | .................... | .................... | .................... |
| N | 4284 | hCoV-19/USA/MA-QDX-1486/2020\|EPI_ISL_603827\|2020-0 | .................... | .................... | .................... | .................... | .................... | .................... |
| N | 4285 | hCoV-19/USA/MA-QDX-1487/2020\|EPI_ISL_603828\|2020-0 | .................... | .................... | .................... | .................... | .................... | .................... |
| N | 4286 | hCoV-19/USA/MA-QDX-1494/2020\|EPI_ISL_603830\|2020-0 | .................... | .................... | .................... | .................... | .................... | .................... |
| N | 4287 | hCoV-19/USA/MA-QDX-1495/2020\|EPI_ISL_603831\|2020-0 | .................... | .................... | .................... | .................... | .................... | .................... |
| N | 4288 | hCoV-19/USA/MA-QDX-1496/2020\|EPI_ISL_603832\|2020-0 | .................... | .................... | .................... | .................... | .................... | .................... |
| N | 4289 | hCoV-19/USA/MA-QDX-1497/2020\|EPI_ISL_603833\|2020-0 | .................... | .................... | .................... | .................... | .................... | .................... |
| N | 4290 | hCoV-19/USA/MA-QDX-1498/2020\|EPI_ISL_603834\|2020-0 | .................... | .................... | .................... | .................... | .................... | .................... |
| N | 4291 | hCoV-19/USA/MA-QDX-1499/2020\|EPI_ISL_603835\|2020-0 | .................... | .................... | .................... | .................... | .................... | .................... |
| N | 4292 | hCoV-19/USA/MA-QDX-1503/2020\|EPI_ISL_603836\|2020-0 | .................... | .................... | .................... | .................... | .................... | .................... |
| N | 4293 | hCoV-19/USA/MA-QDX-1505/2020\|EPI_ISL_603837\|2020-0 | .................... | .................... | .................... | .................... | .................... | .................... |
| N | 4294 | hCoV-19/USA/MA-QDX-1506/2020\|EPI_ISL_603838\|2020-0 | .................... | .................... | .................... | .................... | .................... | .................... |
| N | 4295 | hCoV-19/USA/MA-QDX-1508/2020\|EPI_ISL_603839\|2020-0 | .................... | .................... | .................... | .................... | .................... | .................... |
| N | 4296 | hCoV-19/USA/MA-QDX-1509/2020\|EPI_ISL_603840\|2020-0 | .................... | .................... | .................... | .................... | .................... | .................... |
| N | 4297 | hCoV-19/USA/MI-QDX-1545/2020\|EPI_ISL_603841\|2020-0 | .................... | .................... | .................... | .................... | .................... | .................... |
| N | 4298 | hCoV-19/USA/NY-QDX-1514/2020\|EPI_ISL_603842\|2020-0 | .................... | .................... | .................... | .................... | .................... | .................... |
| N | 4299 | hCoV-19/USA/OH-QDX-1501/2020\|EPI_ISL_603843\|2020-0 | .................... | .................... | .................... | .................... | .................... | .................... |
| N | 4300 | hCoV-19/USA/PA-QDX-1502/2020\|EPI_ISL_603844\|2020-0 | .................... | .................... | .................... | .................... | .................... | .................... |
| N | 4301 | hCoV-19/USA/UT-QDX-1461/2020\|EPI_ISL_603846\|2020-0 | .................... | .................... | .................... | .................... | .................... | .................... |
| N | 4302 | hCoV-19/USA/UT-QDX-1462/2020\|EPI_ISL_603847\|2020-0 | .................... | .................... | .................... | .................... | .................... | .................... |
| N | 4303 | hCoV-19/USA/UT-QDX-1463/2020\|EPI_ISL_603848\|2020-0 | .................... | .................... | .................... | .................... | .................... | .................... |
| N | 4304 | hCoV-19/USA/UT-QDX-1464/2020\|EPI_ISL_603849\|2020-0 | .................... | .................... | .................... | .................... | .................... | .................... |
| N | 4305 | hCoV-19/USA/UT-QDX-1465/2020\|EPI_ISL_603850\|2020-0 | .................... | .................... | .................... | .................... | .................... | .................... |
| N | 4306 | hCoV-19/USA/UT-QDX-1466/2020\|EPI_ISL_603851\|2020-0 | .................... | .................... | .................... | .................... | .................... | .................... |
| N | 4307 | hCoV-19/USA/UT-QDX-1467/2020\|EPI_ISL_603852\|2020-0 | .................... | .................... | .................... | .................... | .................... | .................... |
| N | 4308 | hCoV-19/USA/UT-QDX-1468/2020\|EPI_ISL_603853\|2020-0 | .................... | .................... | .................... | .................... | .................... | .................... |
| N | 4309 | hCoV-19/USA/UT-QDX-1469/2020\|EPI_ISL_603854\|2020-0 | .................... | .................... | .................... | .................... | .................... | .................... |
| N | 4310 | hCoV-19/USA/UT-QDX-1471/2020\|EPI_ISL_603856\|2020-0 | .................... | .................... | .................... | .................... | .................... | .................... |
| N | 4311 | hCoV-19/USA/UT-QDX-1472/2020\|EPI_ISL_603857\|2020-0 | .................... | .................... | .................... | .................... | .................... | .................... |
| N | 4312 | hCoV-19/USA/UT-QDX-1473/2020\|EPI_ISL_603858\|2020-0 | .................... | .................... | .................... | .................... | .................... | .................... |
| N | 4313 | hCoV-19/USA/UT-QDX-1474/2020\|EPI_ISL_603859\|2020-0 | .................... | .................... | .................... | .................... | .................... | .................... |
| N | 4314 | hCoV-19/USA/UT-QDX-1476/2020\|EPI_ISL_603861\|2020-0 | .................... | .................... | .................... | .................... | .................... | .................... |
| N | 4315 | hCoV-19/USA/UT-QDX-1477/2020\|EPI_ISL_603862\|2020-0 | .................... | .................... | .................... | .................... | .................... | .................... |
| N | 4316 | hCoV-19/USA/UT-QDX-1478/2020\|EPI_ISL_603863\|2020-0 | .................... | .................... | .................... | .................... | .................... | .................... |
| N | 4317 | hCoV-19/USA/UT-QDX-1479/2020\|EPI_ISL_603864\|2020-0 | .................... | .................... | .................... | .................... | .................... | .................... |
| N | 4318 | hCoV-19/USA/UT-QDX-1480/2020\|EPI_ISL_603865\|2020-0 | .................... | .................... | .................... | .................... | .................... | .................... |
| N | 4319 | hCoV-19/USA/UT-QDX-1481/2020\|EPI_ISL_603866\|2020-0 | .................... | .................... | .................... | .................... | .................... | .................... |
| N | 4320 | hCoV-19/USA/UT-QDX-1482/2020\|EPI_ISL_603867\|2020-0 | .................... | .................... | .................... | .................... | .................... | .................... |
| N | 4321 | hCoV-19/USA/UT-QDX-1483/2020\|EPI_ISL_603868\|2020-0 | .................... | .................... | .................... | .................... | .................... | .................... |
| N | 4322 | hCoV-19/USA/UT-QDX-1484/2020\|EPI_ISL_603869\|2020-0 | .................... | .................... | .................... | .................... | .................... | .................... |
| N | 4323 | hCoV-19/USA/UT-QDX-1485/2020\|EPI_ISL_603870\|2020-0 | .................... | .................... | .................... | .................... | .................... | .................... |
| N | 4324 | hCoV-19/USA/UT-QDX-1500/2020\|EPI_ISL_603871\|2020-0 | .................... | .................... | .................... | .................... | .................... | .................... |
| N | 4325 | hCoV-19/USA/AZ-QDX-1568/2020\|EPI_ISL_603872\|2020-0 | .................... | .................... | .................... | .................... | .................... | .................... |
| N | 4326 | hCoV-19/USA/AZ-QDX-1580/2020\|EPI_ISL_603873\|2020-0 | .................... | .................... | .................... | .................... | .................... | .................... |
| N | 4327 | hCoV-19/USA/AZ-QDX-1585/2020\|EPI_ISL_603874\|2020-0 | .................... | .................... | .................... | .................... | .................... | .................... |
| N | 4328 | hCoV-19/USA/AZ-QDX-1610/2020\|EPI_ISL_603875\|2020-0 | .................... | .................... | .................... | .................... | .................... | .................... |
| N | 4329 | hCoV-19/USA/AZ-QDX-1626/2020\|EPI_ISL_603876\|2020-0 | .................... | .................... | .................... | .................... | .................... | .................... |
| N | 4330 | hCoV-19/USA/AZ-QDX-1645/2020\|EPI_ISL_603877\|2020-0 | .................... | .................... | .................... | .................... | .................... | .................... |
| N | 4331 | hCoV-19/USA/AZ-QDX-1660/2020\|EPI_ISL_603878\|2020-0 | .................... | .................... | .................... | .................... | .................... | .................... |
| N | 4332 | hCoV-19/USA/AZ-QDX-1667/2020\|EPI_ISL_603879\|2020-0 | .................... | .................... | .................... | .................... | .................... | .................... |
| N | 4333 | hCoV-19/USA/AZ-QDX-1678/2020\|EPI_ISL_603880\|2020-0 | .................... | .................... | .................... | .................... | .................... | .................... |
| N | 4334 | hCoV-19/USA/AZ-QDX-1692/2020\|EPI_ISL_603881\|2020-0 | .................... | .................... | .................... | .................... | .................... | .................... |
| N | 4335 | hCoV-19/USA/AZ-QDX-1699/2020\|EPI_ISL_603882\|2020-0 | .................... | .................... | .................... | .................... | .................... | .................... |
| N | 4336 | hCoV-19/USA/CA-QDX-1559/2020\|EPI_ISL_603883\|2020-0 | .................... | .................... | .................... | .................... | .................... | .................... |
| N | 4337 | hCoV-19/USA/CA-QDX-1560/2020\|EPI_ISL_603884\|2020-0 | .................... | .................... | .................... | .................... | .................... | .................... |
| N | 4338 | hCoV-19/USA/CA-QDX-1562/2020\|EPI_ISL_603885\|2020-0 | .................... | .................... | .................... | .................... | .................... | .................... |
| N | 4339 | hCoV-19/USA/CA-QDX-1566/2020\|EPI_ISL_603886\|2020-0 | .................... | .................... | .................... | .................... | .................... | .................... |
| N | 4340 | hCoV-19/USA/CA-QDX-1567/2020\|EPI_ISL_603887\|2020-0 | .................... | .................... | .................... | .................... | .................... | .................... |
| N | 4341 | hCoV-19/USA/CA-QDX-1571/2020\|EPI_ISL_603888\|2020-0 | .................... | .................... | .................... | .................... | .................... | .................... |
| N | 4342 | hCoV-19/USA/CA-QDX-1572/2020\|EPI_ISL_603889\|2020-0 | .................... | .................... | .................... | .................... | .................... | .................... |
| N | 4343 | hCoV-19/USA/CA-QDX-1576/2020\|EPI_ISL_603890\|2020-0 | .................... | .................... | .................... | .................... | .................... | .................... |

| | | | | | | | | |
|---|---|---|---|---|---|---|---|---|
| N | 4344 | hCoV-19/USA/CA-QDX-1577/2020\|EPI_ISL_603891\|2020-0 | ···················· | ···················· | ···················· | ···················· | ···················· | ···················· |
| N | 4345 | hCoV-19/USA/CA-QDX-1578/2020\|EPI_ISL_603892\|2020-0 | ···················· | ···················· | ···················· | ···················· | ···················· | ···················· |
| N | 4346 | hCoV-19/USA/CA-QDX-1579/2020\|EPI_ISL_603893\|2020-0 | ···················· | ···················· | ···················· | ···················· | ···················· | ···················· |
| N | 4347 | hCoV-19/USA/CA-QDX-1583/2020\|EPI_ISL_603894\|2020-0 | ···················· | ···················· | ···················· | ···················· | ···················· | ···················· |
| N | 4348 | hCoV-19/USA/CA-QDX-1584/2020\|EPI_ISL_603895\|2020-0 | ···················· | ···················· | ···················· | ···················· | ···················· | ···················· |
| N | 4349 | hCoV-19/USA/CA-QDX-1586/2020\|EPI_ISL_603896\|2020-0 | ···················· | ···················· | ···················· | ···················· | ···················· | ···················· |
| N | 4350 | hCoV-19/USA/CA-QDX-1587/2020\|EPI_ISL_603897\|2020-0 | ···················· | ···················· | ···················· | ···················· | ···················· | ···················· |
| N | 4351 | hCoV-19/USA/CA-QDX-1589/2020\|EPI_ISL_603898\|2020-0 | ···················· | ···················· | ···················· | ···················· | ···················· | ···················· |
| N | 4352 | hCoV-19/USA/CA-QDX-1598/2020\|EPI_ISL_603899\|2020-0 | ···················· | ···················· | ···················· | ···················· | ···················· | ···················· |
| N | 4353 | hCoV-19/England/QEUH-A5E449/2020\|EPI_ISL_597511\|20 | ···················· | ·A·······A·········· | ···················· | ···················· | ···················· | ···················· |
| N | 4354 | hCoV-19/England/CAMC-A57A11/2020\|EPI_ISL_597512\|20 | ···················· | ···················· | ···················· | ···················· | ···················· | ···················· |
| N | 4355 | hCoV-19/England/QEUH-A5E458/2020\|EPI_ISL_597513\|20 | ···················· | ···················· | ···················· | ···················· | ···················· | ···················· |
| N | 4356 | hCoV-19/England/QEUH-A5E203/2020\|EPI_ISL_597514\|20 | ···················· | ···················· | ···················· | ···················· | ···················· | ···················· |
| N | 4357 | hCoV-19/England/QEUH-A5E537/2020\|EPI_ISL_597515\|20 | ···················· | ···················· | ···················· | ···················· | ···················· | ···················· |
| N | 4358 | hCoV-19/England/QEUH-A5E27C/2020\|EPI_ISL_597516\|20 | ···················· | ···················· | ···················· | ···················· | ···················· | ···················· |
| N | 4359 | hCoV-19/England/CAMC-A57194/2020\|EPI_ISL_597517\|20 | ···················· | ···················· | ···················· | ···················· | ···················· | ···················· |
| N | 4360 | hCoV-19/England/QEUH-A5DDC7/2020\|EPI_ISL_597518\|20 | ···················· | ···················· | ···················· | ···················· | ···················· | ···················· |
| N | 4361 | hCoV-19/England/CAMC-A570C4/2020\|EPI_ISL_597519\|20 | ···················· | ···················· | ···················· | ···················· | ···················· | ···················· |
| N | 4362 | hCoV-19/England/CAMC-A572FB/2020\|EPI_ISL_597520\|20 | ···················· | ···················· | ···················· | ···················· | ···················· | ···················· |
| N | 4363 | hCoV-19/England/QEUH-A5E24F/2020\|EPI_ISL_597521\|20 | ···················· | ···················· | ···················· | ···················· | ···················· | ···················· |
| N | 4364 | hCoV-19/England/ALDP-A504B0/2020\|EPI_ISL_597523\|20 | ···················· | ···················· | ···················· | ···················· | ···················· | ···················· |
| N | 4365 | hCoV-19/England/ALDP-A50386/2020\|EPI_ISL_597524\|20 | ···················· | ···················· | ···················· | ···················· | ···················· | ···················· |
| N | 4366 | hCoV-19/England/ALDP-A509FD/2020\|EPI_ISL_597525\|20 | ···················· | ·A·················· | ···················· | ···················· | ···················· | ···A················ |
| N | 4367 | hCoV-19/England/ALDP-A50E2B/2020\|EPI_ISL_597526\|20 | ···················· | ···················· | ···················· | ···················· | ···················· | ···················· |
| N | 4368 | hCoV-19/England/ALDP-A50896/2020\|EPI_ISL_597527\|20 | ···················· | ···················· | ···················· | ···················· | ···················· | ···················· |
| N | 4369 | hCoV-19/England/ALDP-A51051/2020\|EPI_ISL_597529\|20 | ···················· | ···················· | ···················· | ···················· | ···················· | ···················· |
| N | 4370 | hCoV-19/England/ALDP-A50DF1/2020\|EPI_ISL_597530\|20 | ···················· | ···················· | ···················· | ···················· | ···················· | ···················· |
| N | 4371 | hCoV-19/England/ALDP-A4FC8F/2020\|EPI_ISL_597531\|20 | ···················· | ···················· | ···················· | ···················· | ···················· | ···················· |
| N | 4372 | hCoV-19/England/ALDP-A510D9/2020\|EPI_ISL_597532\|20 | ···················· | ···················· | ···················· | ···················· | ···················· | ···················· |
| N | 4373 | hCoV-19/England/ALDP-A4FB64/2020\|EPI_ISL_597533\|20 | ···················· | ···················· | ···················· | ···················· | ···················· | ···················· |
| N | 4374 | hCoV-19/England/ALDP-A50562/2020\|EPI_ISL_597534\|20 | ···················· | ···················· | ···················· | ···················· | ···················· | ···················· |
| N | 4375 | hCoV-19/England/ALDP-A50D00/2020\|EPI_ISL_597535\|20 | ···················· | ···················· | ···················· | ···················· | ···················· | ···················· |
| N | 4376 | hCoV-19/England/ALDP-A50AFA/2020\|EPI_ISL_597536\|20 | ···················· | ···················· | ···················· | ···················· | ···················· | ···················· |
| N | 4377 | hCoV-19/England/ALDP-A509A2/2020\|EPI_ISL_597537\|20 | ···················· | ···················· | ···················· | ···················· | ···················· | ···················· |
| N | 4378 | hCoV-19/England/ALDP-A507D5/2020\|EPI_ISL_597538\|20 | ···················· | ···················· | ···················· | ···················· | ···················· | ···················· |
| N | 4379 | hCoV-19/England/ALDP-A5068D/2020\|EPI_ISL_597539\|20 | ···················· | ···················· | ···················· | ···················· | ···················· | ···················· |
| N | 4380 | hCoV-19/England/ALDP-A507A8/2020\|EPI_ISL_597540\|20 | ···················· | ···················· | ···················· | ···················· | ···················· | ···················· |
| N | 4381 | hCoV-19/England/ALDP-A50210/2020\|EPI_ISL_597542\|20 | ···················· | ···················· | ···················· | ···················· | ···················· | ···················· |
| N | 4382 | hCoV-19/England/ALDP-A50F0A/2020\|EPI_ISL_597543\|20 | ···················· | ···················· | ···················· | ···················· | ···················· | ···················· |
| N | 4383 | hCoV-19/England/ALDP-A510CA/2020\|EPI_ISL_597544\|20 | ···················· | ···················· | ···················· | ···················· | ···················· | ···················· |
| N | 4384 | hCoV-19/England/ALDP-A4FCBC/2020\|EPI_ISL_597545\|20 | ···················· | ···················· | ···················· | ···················· | ···················· | ···················· |
| N | 4385 | hCoV-19/England/ALDP-A50B42/2020\|EPI_ISL_597546\|20 | ···················· | ···················· | ···················· | ···················· | ···················· | ···················· |
| N | 4386 | hCoV-19/England/ALDP-A50131/2020\|EPI_ISL_597547\|20 | ···················· | ···················· | ···················· | ···················· | ···················· | ···················· |
| N | 4387 | hCoV-19/England/ALDP-A4FCDA/2020\|EPI_ISL_597548\|20 | ···················· | ···················· | ···················· | ···················· | ···················· | ···················· |
| N | 4388 | hCoV-19/England/ALDP-A4FE89/2020\|EPI_ISL_597549\|20 | ···················· | ···················· | ···················· | ···················· | ···················· | ···················· |
| N | 4389 | hCoV-19/England/ALDP-A4FCAD/2020\|EPI_ISL_597550\|20 | ···················· | ···················· | ···················· | ···················· | ···················· | ···················· |
| N | 4390 | hCoV-19/England/ALDP-A50429/2020\|EPI_ISL_597551\|20 | ···················· | ···················· | ···················· | ···················· | ···················· | ···················· |
| N | 4391 | hCoV-19/England/ALDP-A50FEC/2020\|EPI_ISL_597552\|20 | ···················· | ···················· | ···················· | ···················· | ···················· | ···················· |
| N | 4392 | hCoV-19/England/ALDP-A4FFB3/2020\|EPI_ISL_597553\|20 | ···················· | ···················· | ···················· | ···················· | ···················· | ···················· |
| N | 4393 | hCoV-19/England/ALDP-A509DF/2020\|EPI_ISL_597554\|20 | ···················· | ···················· | ···················· | ···················· | ···················· | ···················· |
| N | 4394 | hCoV-19/England/MILK-A44CD5/2020\|EPI_ISL_598810\|20 | ···················· | ···················· | ···················· | ···················· | ···················· | ···················· |
| N | 4395 | hCoV-19/England/MILK-A4571F/2020\|EPI_ISL_598811\|20 | ···················· | ···················· | ···················· | ···················· | ···················· | ···················· |
| N | 4396 | hCoV-19/England/MILK-A4631A/2020\|EPI_ISL_598812\|20 | ···········T········ | ···················· | ···················· | ···················· | ···················· | ···················· |
| N | 4397 | hCoV-19/England/MILK-A460B9/2020\|EPI_ISL_598813\|20 | ···················· | ···················· | ···················· | ···················· | ···················· | ···················· |
| N | 4398 | hCoV-19/England/MILK-A4609B/2020\|EPI_ISL_598816\|20 | ···················· | ···················· | ···················· | ···················· | ···················· | ···················· |
| N | 4399 | hCoV-19/Scotland/QEUH-A2197D/2020\|EPI_ISL_598842\|2 | ···················· | ···················· | ···················· | ···················· | ···················· | ···················· |
| N | 4400 | hCoV-19/Scotland/QEUH-A21940/2020\|EPI_ISL_598843\|2 | ···················· | ···················· | ···················· | ···················· | ···················· | ···················· |
| N | 4401 | hCoV-19/Scotland/QEUH-A1E3C0/2020\|EPI_ISL_598844\|2 | ···················· | ···················· | ···················· | ···················· | ···················· | ···················· |
| N | 4402 | hCoV-19/Scotland/QEUH-A21807/2020\|EPI_ISL_598845\|2 | ···················· | ···················· | ···················· | ···················· | ···················· | ···················· |
| N | 4403 | hCoV-19/Scotland/QEUH-A1E3FD/2020\|EPI_ISL_598846\|2 | ···················· | ···················· | ···················· | ···················· | ···················· | ···················· |
| N | 4404 | hCoV-19/Scotland/QEUH-A1E050/2020\|EPI_ISL_598847\|2 | ···················· | ···················· | ···················· | ···················· | ···················· | ···················· |
| N | 4405 | hCoV-19/Scotland/QEUH-A1E409/2020\|EPI_ISL_598849\|2 | ···················· | ···················· | ···················· | ···················· | ···················· | ···················· |
| N | 4406 | hCoV-19/Scotland/QEUH-A1DA17/2020\|EPI_ISL_598850\|2 | ···················· | ···················· | ···················· | ···················· | ···················· | ···················· |
| N | 4407 | hCoV-19/Scotland/QEUH-A1EA07/2020\|EPI_ISL_598851\|2 | ···················· | ···················· | ···················· | ···················· | ···················· | ···················· |
| N | 4408 | hCoV-19/Scotland/QEUH-A219AA/2020\|EPI_ISL_598852\|2 | ···················· | ···················· | ···················· | ···················· | ···················· | ···················· |
| N | 4409 | hCoV-19/Scotland/QEUH-A214F4/2020\|EPI_ISL_598853\|2 | ···················· | ···················· | ···················· | ···················· | ···················· | ···················· |
| N | 4410 | hCoV-19/Scotland/QEUH-A1EE74/2020\|EPI_ISL_598854\|2 | ···················· | ···················· | ···················· | ···················· | ···················· | ···················· |
| N | 4411 | hCoV-19/Scotland/QEUH-A1EA61/2020\|EPI_ISL_598855\|2 | ···················· | ···················· | ···················· | ···················· | ···················· | ···················· |
| N | 4412 | hCoV-19/Scotland/QEUH-A1E0F6/2020\|EPI_ISL_598856\|2 | ···················· | ···················· | ···················· | ···················· | ···················· | ···················· |
| N | 4413 | hCoV-19/Scotland/QEUH-A1EA16/2020\|EPI_ISL_598857\|2 | ···················· | ···················· | ···················· | ···················· | ···················· | ···················· |
| N | 4414 | hCoV-19/Scotland/QEUH-A1DA08/2020\|EPI_ISL_598858\|2 | ···················· | ···················· | ···················· | ···················· | ···················· | ···················· |
| N | 4415 | hCoV-19/Scotland/QEUH-A2199B/2020\|EPI_ISL_598859\|2 | ···················· | ···················· | ···················· | ···················· | ···················· | ···················· |
| N | 4416 | hCoV-19/Scotland/QEUH-A1E0D8/2020\|EPI_ISL_598860\|2 | ···················· | ···················· | ···················· | ···················· | ···················· | ···················· |
| N | 4417 | hCoV-19/Scotland/QEUH-A1EDD1/2020\|EPI_ISL_598861\|2 | ···················· | ···················· | ···················· | ···················· | ···················· | ···················· |
| N | 4418 | hCoV-19/Scotland/QEUH-A21816/2020\|EPI_ISL_598862\|2 | ···················· | ···················· | ···················· | ···················· | ···················· | ···················· |
| N | 4419 | hCoV-19/Scotland/QEUH-A21931/2020\|EPI_ISL_598863\|2 | ···················· | ···················· | ···················· | ···················· | ···················· | ···················· |
| N | 4420 | hCoV-19/Scotland/QEUH-A21F8A/2020\|EPI_ISL_598864\|2 | ···················· | ···················· | ···················· | ···················· | ···················· | ···················· |
| N | 4421 | hCoV-19/Scotland/QEUH-A1E506/2020\|EPI_ISL_598865\|2 | ···················· | ···················· | ···················· | ···················· | ···················· | ···················· |
| N | 4422 | hCoV-19/Scotland/QEUH-A1E120/2020\|EPI_ISL_598866\|2 | ···················· | ···················· | ···················· | ···················· | ···················· | ···················· |
| N | 4423 | hCoV-19/Scotland/QEUH-A21922/2020\|EPI_ISL_598867\|2 | ···················· | ···················· | ···················· | ···················· | ···················· | ···················· |

| | | | | | | | | |
|---|---|---|---|---|---|---|---|---|
| N | 4424 | hCoV-19/Scotland/QEUH-A1E700/2020\|EPI_ISL_598868\|2 | ···················· | ·A·················· | ···················· | ···················· | ···················· | ···················· |
| N | 4425 | hCoV-19/Scotland/QEUH-A1E427/2020\|EPI_ISL_598869\|2 | ···················· | ···················· | ···················· | ···················· | ···················· | ···················· |
| N | 4426 | hCoV-19/Scotland/QEUH-A1E9FB/2020\|EPI_ISL_598870\|2 | ···················· | ···················· | ···················· | ···················· | ···················· | ···················· |
| N | 4427 | hCoV-19/Scotland/QEUH-A2155B/2020\|EPI_ISL_598871\|2 | ···················· | ···················· | ···················· | ···················· | ···················· | ···················· |
| N | 4428 | hCoV-19/Scotland/QEUH-A2154C/2020\|EPI_ISL_598872\|2 | ···················· | ···················· | ···················· | ···················· | ···················· | ···················· |
| N | 4429 | hCoV-19/Scotland/QEUH-A1E542/2020\|EPI_ISL_598873\|2 | ···················· | ···················· | ···················· | ···················· | ···················· | ···················· |
| N | 4430 | hCoV-19/Scotland/QEUH-A21649/2020\|EPI_ISL_598874\|2 | ···················· | ···················· | ···················· | ···················· | ···················· | ···················· |
| N | 4431 | hCoV-19/Scotland/QEUH-A218CB/2020\|EPI_ISL_598876\|2 | ···················· | ···················· | ···················· | ···················· | ···················· | ···················· |
| N | 4432 | hCoV-19/Scotland/QEUH-A1E6F4/2020\|EPI_ISL_598877\|2 | ···················· | ···················· | ···················· | ···················· | ···················· | ···················· |
| N | 4433 | hCoV-19/Scotland/QEUH-A1E0C9/2020\|EPI_ISL_598878\|2 | ···················· | ···················· | ···················· | ···················· | ···················· | ···················· |
| N | 4434 | hCoV-19/Scotland/QEUH-A1E58E/2020\|EPI_ISL_598879\|2 | ···················· | ···················· | ···················· | ···················· | ···················· | ···················· |
| N | 4435 | hCoV-19/Scotland/QEUH-A1E0BA/2020\|EPI_ISL_598880\|2 | ···················· | ···················· | ···················· | ···················· | ···················· | ···················· |
| N | 4436 | hCoV-19/Scotland/QEUH-A1E533/2020\|EPI_ISL_598881\|2 | ···················· | ···················· | ···················· | ···················· | ···················· | ···················· |
| N | 4437 | hCoV-19/Scotland/QEUH-A1EA34/2020\|EPI_ISL_598882\|2 | ···················· | ···················· | ···················· | ···················· | ···················· | ···················· |
| N | 4438 | hCoV-19/Scotland/QEUH-A1E418/2020\|EPI_ISL_598883\|2 | ···················· | ···················· | ···················· | ···················· | ···················· | ···················· |
| N | 4439 | hCoV-19/Scotland/QEUH-A1E5CA/2020\|EPI_ISL_598884\|2 | ···················· | ···················· | ···················· | ···················· | ···················· | ···················· |
| N | 4440 | hCoV-19/Scotland/QEUH-A21579/2020\|EPI_ISL_598885\|2 | ···················· | ···················· | ···················· | ···················· | ···················· | ···················· |
| N | 4441 | hCoV-19/Scotland/QEUH-9DC426/2020\|EPI_ISL_601100\|2 | ···················· | ···················· | ···················· | ···················· | ···················· | ···················· |
| N | 4442 | hCoV-19/Scotland/QEUH-9DC3A1/2020\|EPI_ISL_601101\|2 | ···················· | ···················· | ···················· | ···················· | ···················· | ···················· |
| N | 4443 | hCoV-19/Scotland/QEUH-9DC4CC/2020\|EPI_ISL_601102\|2 | ···················· | ···················· | ···················· | ···················· | ···················· | ···················· |
| N | 4444 | hCoV-19/Scotland/QEUH-9DAD69/2020\|EPI_ISL_601103\|2 | ···················· | ···················· | ···················· | ···················· | ···················· | ···················· |
| N | 4445 | hCoV-19/Scotland/QEUH-9DAF18/2020\|EPI_ISL_601104\|2 | ···················· | ···················· | ···················· | ···················· | ···················· | ···················· |
| N | 4446 | hCoV-19/Scotland/QEUH-9DAF45/2020\|EPI_ISL_601105\|2 | ···················· | ···················· | ···················· | ···················· | ···················· | ···················· |
| N | 4447 | hCoV-19/Scotland/QEUH-9DCAE8/2020\|EPI_ISL_601106\|2 | ···················· | ···················· | ···················· | ···················· | ···················· | ···················· |
| N | 4448 | hCoV-19/Scotland/QEUH-9DAD78/2020\|EPI_ISL_601107\|2 | ···················· | ·A·················· | ···················· | ···················· | ···················· | ···················· |
| N | 4449 | hCoV-19/Scotland/QEUH-9DCACA/2020\|EPI_ISL_601108\|2 | ···················· | ···················· | ···················· | ···················· | ···················· | ···················· |
| N | 4450 | hCoV-19/Scotland/QEUH-9DC17A/2020\|EPI_ISL_601109\|2 | ···················· | ···················· | ···················· | ···················· | ···················· | ···················· |
| N | 4451 | hCoV-19/Scotland/QEUH-9DC514/2020\|EPI_ISL_601130\|2 | ···················· | ···················· | ···················· | ···················· | ···················· | ···················· |
| N | 4452 | hCoV-19/Scotland/QEUH-9DACB7/2020\|EPI_ISL_601131\|2 | ···················· | ···················· | ···················· | ···················· | ···················· | ···················· |
| N | 4453 | hCoV-19/Scotland/QEUH-9DCEA0/2020\|EPI_ISL_601132\|2 | ···················· | ···················· | ···················· | ···················· | ···················· | ···················· |
| N | 4454 | hCoV-19/Scotland/QEUH-9DAC7B/2020\|EPI_ISL_601133\|2 | ···················· | ···················· | ···················· | ···················· | ···················· | ···················· |
| N | 4455 | hCoV-19/Scotland/QEUH-9DAE48/2020\|EPI_ISL_601134\|2 | ···················· | ···················· | ···················· | ···················· | ···················· | ···················· |
| N | 4456 | hCoV-19/Scotland/QEUH-9DC329/2020\|EPI_ISL_601135\|2 | ···················· | ···················· | ···················· | ···················· | ···················· | ···················· |
| N | 4457 | hCoV-19/Scotland/QEUH-9DAB6F/2020\|EPI_ISL_601136\|2 | ···················· | ···················· | ···················· | ···················· | ···················· | ···················· |
| N | 4458 | hCoV-19/Scotland/QEUH-9DAEB1/2020\|EPI_ISL_601137\|2 | ···················· | ···················· | ···················· | ···················· | ···················· | ···················· |
| N | 4459 | hCoV-19/Scotland/QEUH-9DABF6/2020\|EPI_ISL_601138\|2 | ···················· | ···················· | ···················· | ···················· | ···················· | ···················· |
| N | 4460 | hCoV-19/Scotland/QEUH-9DC49F/2020\|EPI_ISL_601139\|2 | ···················· | ···················· | ···················· | ···················· | ···················· | ···················· |
| N | 4461 | hCoV-19/Scotland/QEUH-9DC505/2020\|EPI_ISL_601140\|2 | ···················· | ···················· | ···················· | ···················· | ···················· | ···················· |
| N | 4462 | hCoV-19/Scotland/QEUH-9DCE91/2020\|EPI_ISL_601141\|2 | ···················· | ···················· | ···················· | ···················· | ···················· | ···················· |
| N | 4463 | hCoV-19/Scotland/QEUH-9DAF90/2020\|EPI_ISL_601142\|2 | ···················· | ···················· | ···················· | ···················· | ···················· | ···················· |
| N | 4464 | hCoV-19/Scotland/QEUH-9DAC02/2020\|EPI_ISL_601143\|2 | ···················· | ···················· | ···················· | ···················· | ···················· | ···················· |
| N | 4465 | hCoV-19/Scotland/QEUH-9DAB41/2020\|EPI_ISL_601144\|2 | ···················· | ···················· | ···················· | ···················· | ···················· | ···················· |
| N | 4466 | hCoV-19/Scotland/QEUH-9DC4EA/2020\|EPI_ISL_601145\|2 | ···················· | ···················· | ···················· | ···················· | ···················· | ···················· |
| N | 4467 | hCoV-19/Scotland/QEUH-9DCEBF/2020\|EPI_ISL_601146\|2 | ···················· | ···················· | ···················· | ···················· | ···················· | ···················· |
| N | 4468 | hCoV-19/Scotland/QEUH-9DACA8/2020\|EPI_ISL_601147\|2 | ···················· | ···················· | ···················· | ···················· | ···················· | ···················· |
| N | 4469 | hCoV-19/Scotland/QEUH-9DAFEB/2020\|EPI_ISL_601148\|2 | ···················· | ···················· | ···················· | ···················· | ···················· | ···················· |
| N | 4470 | hCoV-19/Scotland/QEUH-9DAD87/2020\|EPI_ISL_601149\|2 | ···················· | ···················· | ···················· | ···················· | ······T············· | ···················· |
| N | 4471 | hCoV-19/Scotland/QEUH-9DC4AE/2020\|EPI_ISL_601151\|2 | ···················· | ···················· | ···················· | ···················· | ······T············· | ···················· |
| N | 4472 | hCoV-19/Scotland/QEUH-9DAC99/2020\|EPI_ISL_601152\|2 | ···················· | ···················· | ···················· | ···················· | ···················· | ···················· |
| N | 4473 | hCoV-19/Scotland/QEUH-9DC338/2020\|EPI_ISL_601153\|2 | ···················· | ···················· | ···················· | ···················· | ···················· | ···················· |
| N | 4474 | hCoV-19/Scotland/QEUH-9DC3ED/2020\|EPI_ISL_601154\|2 | ···················· | ···················· | ···················· | ···················· | ···················· | ···················· |
| N | 4475 | hCoV-19/Scotland/QEUH-9DC444/2020\|EPI_ISL_601155\|2 | ···················· | ···················· | ···················· | ···················· | ···················· | ···················· |
| N | 4476 | hCoV-19/Scotland/QEUH-9DAE93/2020\|EPI_ISL_601156\|2 | ···················· | ···················· | ···················· | ···················· | ···················· | ···················· |
| N | 4477 | hCoV-19/Scotland/QEUH-9DC471/2020\|EPI_ISL_601157\|2 | ···················· | ···················· | ···················· | ···················· | ···················· | ···················· |
| N | 4478 | hCoV-19/Scotland/QEUH-9DCB21/2020\|EPI_ISL_601158\|2 | ···················· | ···················· | ···················· | ···················· | ···················· | ···················· |
| N | 4479 | hCoV-19/Scotland/QEUH-9DB014/2020\|EPI_ISL_601159\|2 | ···················· | ···················· | ···················· | ···················· | ···················· | ···················· |
| N | 4480 | hCoV-19/Scotland/QEUH-9DC3DE/2020\|EPI_ISL_601160\|2 | ···················· | ···················· | ···················· | ···················· | ···················· | ···················· |
| N | 4481 | hCoV-19/Scotland/QEUH-9DC408/2020\|EPI_ISL_601161\|2 | ···················· | ···················· | ···················· | ···················· | ···················· | ···················· |
| N | 4482 | hCoV-19/Scotland/QEUH-9DCA15/2020\|EPI_ISL_601162\|2 | ···················· | ···················· | ···················· | ···················· | ···················· | ···················· |
| N | 4483 | hCoV-19/Scotland/QEUH-9DCAF7/2020\|EPI_ISL_601163\|2 | ···················· | ···················· | ···················· | ···················· | ···················· | ···················· |
| N | 4484 | hCoV-19/Scotland/QEUH-9DC3CF/2020\|EPI_ISL_601164\|2 | ···················· | ···················· | ···················· | ···················· | ···················· | ···················· |
| N | 4485 | hCoV-19/Scotland/QEUH-9DCDEF/2020\|EPI_ISL_601165\|2 | ···················· | ···················· | ···················· | ···················· | ···················· | ···················· |
| N | 4486 | hCoV-19/Scotland/QEUH-9DC480/2020\|EPI_ISL_601166\|2 | ···················· | ···················· | ···················· | ···················· | ···················· | ···················· |
| N | 4487 | hCoV-19/Scotland/QEUH-9DAB50/2020\|EPI_ISL_601167\|2 | ···················· | ···················· | ···················· | ···················· | ···················· | ···················· |
| N | 4488 | hCoV-19/Scotland/QEUH-9DC4BD/2020\|EPI_ISL_601168\|2 | ···················· | ···················· | ···················· | ···················· | ···················· | ···················· |
| N | 4489 | hCoV-19/Scotland/QEUH-9DAB14/2020\|EPI_ISL_601169\|2 | ···················· | ···················· | ···················· | ···················· | ···················· | ···················· |
| N | 4490 | hCoV-19/Scotland/QEUH-9DE9F8/2020\|EPI_ISL_601203\|2 | ···················· | ···················· | ···················· | ···················· | ···················· | ···················· |
| N | 4491 | hCoV-19/Russia/AD-RII-MH2738S/2020\|EPI_ISL_602417\| | ···················· | ···················· | ···················· | ···················· | ···················· | ···················· |
| N | 4492 | hCoV-19/Russia/DA-RII-MH2739S/2020\|EPI_ISL_602418\| | ···················· | ···················· | ···················· | ···················· | ···················· | ···················· |
| N | 4493 | hCoV-19/Russia/DA-RII-MH2741S/2020\|EPI_ISL_602419\| | ···················· | ···················· | ···················· | ···················· | ···················· | ···················· |
| N | 4494 | hCoV-19/Russia/DA-RII-MH2742S/2020\|EPI_ISL_602420\| | ···················· | ···················· | ···················· | ···················· | ···················· | ···················· |
| N | 4495 | hCoV-19/Russia/DA-RII-MH2743S/2020\|EPI_ISL_602421\| | ···················· | ···················· | ···················· | ···················· | ···················· | ···················· |
| N | 4496 | hCoV-19/Russia/DA-RII-MH2744S/2020\|EPI_ISL_602422\| | ···················· | ···················· | ···················· | ···················· | ···················· | ···················· |
| N | 4497 | hCoV-19/Russia/IN-RII-MH2745S/2020\|EPI_ISL_602423\| | ···················· | ···················· | ···················· | ···················· | ···················· | ···················· |
| N | 4498 | hCoV-19/Russia/KC-RII-MH2746S/2020\|EPI_ISL_602424\| | ···················· | ···················· | ···················· | ···················· | ···················· | ···················· |
| N | 4499 | hCoV-19/Russia/KC-RII-MH2749S/2020\|EPI_ISL_602425\| | ···················· | ···················· | ···················· | ···················· | ···················· | ···················· |
| N | 4500 | hCoV-19/Russia/KC-RII-MH2751S/2020\|EPI_ISL_602426\| | ···················· | ···················· | ···················· | ···················· | ···················· | ···················· |
| N | 4501 | hCoV-19/Russia/KC-RII-MH2752S/2020\|EPI_ISL_602427\| | ···················· | ···················· | ···················· | ···················· | ···················· | ···················· |
| N | 4502 | hCoV-19/Russia/KC-RII-MH2754S/2020\|EPI_ISL_602428\| | ···················· | ···················· | ···················· | ···················· | ···················· | ···················· |
| N | 4503 | hCoV-19/Russia/KO-RII-MH2755S/2020\|EPI_ISL_602429\| | ···················· | ···················· | ···················· | ···················· | ···················· | ···················· |

| | | | | | | | | |
|---|---|---|---|---|---|---|---|---|
| N | 4504 | hCoV-19/Russia/ME-RII-MH2757S/2020\|EPI_ISL_602430\| | ···················· | ···················· | ····················· | ···················· | ···················· | ···················· |
| N | 4505 | hCoV-19/Russia/KK-RII-MH2758S/2020\|EPI_ISL_602431\| | ···················· | ···················· | ····················· | ···················· | ···················· | ···················· |
| N | 4506 | hCoV-19/Russia/KK-RII-MH2759S/2020\|EPI_ISL_602432\| | ···················· | ···················· | ····················· | ···················· | ···················· | ···················· |
| N | 4507 | hCoV-19/Russia/KK-RII-MH2760S/2020\|EPI_ISL_602433\| | ···················· | ···················· | ····················· | ···················· | ···················· | ···················· |
| N | 4508 | hCoV-19/Russia/ALT-RII-MH2761S/2020\|EPI_ISL_602434 | ···················· | ···················· | ····················· | ···················· | ···················· | ···················· |
| N | 4509 | hCoV-19/Russia/ALT-RII-MH2762S/2020\|EPI_ISL_602435 | ···················· | ···················· | ····················· | ···················· | ···················· | ···················· |
| N | 4510 | hCoV-19/Russia/ALT-RII-MH2764S/2020\|EPI_ISL_602436 | ···················· | ···················· | ····················· | ···················· | ···················· | ···················· |
| N | 4511 | hCoV-19/Russia/ALT-RII-MH2765S/2020\|EPI_ISL_602437 | ···················· | ···················· | ····················· | ···················· | ···················· | ···················· |
| N | 4512 | hCoV-19/Russia/KDA-RII-MH2766S/2020\|EPI_ISL_602438 | ···················· | ···················· | ····················· | ···················· | ···················· | ···················· |
| N | 4513 | hCoV-19/Russia/KDA-RII-MH2768S/2020\|EPI_ISL_602439 | ···················· | ···················· | ····················· | ···················· | ···················· | ···················· |
| N | 4514 | hCoV-19/Russia/MOS-RII-MH2798S/2020\|EPI_ISL_602460 | ···················· | ···················· | ····················· | ···················· | ···················· | ···················· |
| N | 4515 | hCoV-19/Russia/MOS-RII-MH2799S/2020\|EPI_ISL_602461 | ···················· | ···················· | ····················· | ···················· | ···················· | ···················· |
| N | 4516 | hCoV-19/Germany/NW-HHU-189/2020\|EPI_ISL_602463\|202 | ···················· | ···················· | ····················· | ···················· | ···················· | ···················· |
| N | 4517 | hCoV-19/Germany/NW-HHU-190/2020\|EPI_ISL_602464\|202 | ···················· | ···················· | ····················· | ···················· | ···················· | ···················· |
| N | 4518 | hCoV-19/Germany/NW-HHU-191/2020\|EPI_ISL_602465\|202 | ···················· | ···················· | ····················· | ···················· | ···················· | ···················· |
| N | 4520 | hCoV-19/Germany/NW-HHU-193/2020\|EPI_ISL_602467\|202 | ···················· | ···················· | ····················· | ···················· | ···················· | ···················· |
| N | 4521 | hCoV-19/Germany/NW-HHU-194/2020\|EPI_ISL_602468\|202 | ···················· | ···················· | ····················· | ···················· | ···················· | ···················· |
| N | 4522 | hCoV-19/Germany/NW-HHU-195/2020\|EPI_ISL_602469\|202 | ···················· | ···················· | ····················· | ···················· | ···················· | ···················· |
| N | 4523 | hCoV-19/Germany/NW-HHU-196/2020\|EPI_ISL_602470\|202 | ···················· | ···················· | ····················· | ···················· | ···················· | ···················· |
| N | 4524 | hCoV-19/Germany/NW-HHU-197/2020\|EPI_ISL_602471\|202 | ···················· | ···················· | ····················· | ···················· | ···················· | ···················· |
| N | 4525 | hCoV-19/Germany/NW-HHU-198/2020\|EPI_ISL_602472\|202 | ···················· | ···················· | ····················· | ···················· | ···················· | ···················· |
| N | 4526 | hCoV-19/Germany/NW-HHU-199/2020\|EPI_ISL_602473\|202 | ···················· | ···················· | ····················· | ···················· | ···················· | ···················· |
| N | 4527 | hCoV-19/Germany/NW-HHU-200/2020\|EPI_ISL_602474\|202 | ···················· | ···················· | ····················· | ···················· | ···················· | ···················· |
| N | 4528 | hCoV-19/Germany/NW-HHU-201/2020\|EPI_ISL_602475\|202 | ···················· | ···················· | ····················· | ···················· | ···················· | ···················· |
| N | 4529 | hCoV-19/Germany/NW-HHU-202/2020\|EPI_ISL_602476\|202 | ···················· | ···················· | ····················· | ···················· | ···················· | ···················· |
| N | 4530 | hCoV-19/Germany/NW-HHU-203/2020\|EPI_ISL_602477\|202 | ···················· | ···················· | ····················· | ···················· | ···················· | ···················· |
| N | 4531 | hCoV-19/Germany/NW-HHU-204/2020\|EPI_ISL_602478\|202 | ···················· | ···················· | ····················· | ···················· | ···················· | ···················· |
| N | 4532 | hCoV-19/Germany/NW-HHU-205/2020\|EPI_ISL_602479\|202 | ···················· | ···················· | ····················· | ···················· | ···················· | ···················· |
| N | 4533 | hCoV-19/Germany/NW-HHU-206/2020\|EPI_ISL_602480\|202 | ···················· | ···················· | ····················· | ···················· | ···················· | ···················· |
| N | 4534 | hCoV-19/Germany/NW-HHU-207/2020\|EPI_ISL_602481\|202 | ···················· | ···················· | ····················· | ···················· | ···················· | ···················· |
| N | 4535 | hCoV-19/Germany/NW-HHU-208/2020\|EPI_ISL_602482\|202 | ···················· | ···················· | ····················· | ···················· | ···················· | ···················· |
| N | 4536 | hCoV-19/Germany/NW-HHU-209/2020\|EPI_ISL_602483\|202 | ···················· | ···················· | ····················· | ···················· | ···················· | ···················· |
| N | 4537 | hCoV-19/Germany/NW-HHU-210/2020\|EPI_ISL_602484\|202 | ···················· | ···················· | ····················· | ···················· | ···················· | ···················· |
| N | 4538 | hCoV-19/Germany/NW-HHU-211/2020\|EPI_ISL_602485\|202 | ···················· | ···················· | ····················· | ···················· | ···················· | ···················· |
| N | 4539 | hCoV-19/Germany/NW-HHU-165/2020\|EPI_ISL_602487\|202 | ···················· | ···················· | ····················· | ···················· | ···················· | ···················· |
| N | 4540 | hCoV-19/Germany/NW-HHU-166/2020\|EPI_ISL_602488\|202 | ···················· | ···················· | ····················· | ···················· | ···················· | ···················· |
| N | 4541 | hCoV-19/Germany/NW-HHU-167/2020\|EPI_ISL_602489\|202 | ···················· | ···················· | ····················· | ···················· | ···················· | ···················· |
| N | 4543 | hCoV-19/Germany/NW-HHU-169/2020\|EPI_ISL_602491\|202 | ···················· | ···················· | ····················· | ···················· | ···················· | ···················· |
| N | 4544 | hCoV-19/Germany/NW-HHU-170/2020\|EPI_ISL_602492\|202 | ···················· | ···················· | ····················· | ···················· | ···················· | ···················· |
| N | 4545 | hCoV-19/Germany/NW-HHU-171/2020\|EPI_ISL_602493\|202 | ···················· | ···················· | ····················· | ···················· | ···················· | ···················· |
| N | 4546 | hCoV-19/Germany/NW-HHU-172/2020\|EPI_ISL_602494\|202 | ···················· | ···················· | ····················· | ···················· | ···················· | ···················· |
| N | 4547 | hCoV-19/Germany/NW-HHU-173/2020\|EPI_ISL_602495\|202 | ···················· | ···················· | ····················· | ···················· | ···················· | ···················· |
| N | 4548 | hCoV-19/Germany/NW-HHU-174/2020\|EPI_ISL_602496\|202 | ···················· | ···················· | ····················· | ···················· | ···················· | ···················· |
| N | 4549 | hCoV-19/Germany/NW-HHU-175/2020\|EPI_ISL_602497\|202 | ···················· | ···················· | ····················· | ···················· | ···················· | ···················· |
| N | 4550 | hCoV-19/Germany/NW-HHU-176/2020\|EPI_ISL_602498\|202 | ···················· | ···················· | ····················· | ···················· | ···················· | ···················· |
| N | 4551 | hCoV-19/Germany/NW-HHU-177/2020\|EPI_ISL_602499\|202 | ···················· | ···················· | ····················· | ···················· | ···················· | ···················· |
| N | 4552 | hCoV-19/Switzerland/UR-ETHZ-321539/2020\|EPI_ISL_60 | ···················· | ···················· | ····················· | ···················· | ···················· | ···················· |
| N | 4553 | hCoV-19/Switzerland/VD-ETHZ-321540/2020\|EPI_ISL_60 | ···················· | ···················· | ····················· | ···················· | ···················· | ···················· |
| N | 4554 | hCoV-19/Switzerland/UR-ETHZ-321541/2020\|EPI_ISL_60 | ···················· | ···················· | ····················· | ···················· | ···················· | ···················· |
| N | 4555 | hCoV-19/Switzerland/UR-ETHZ-321542/2020\|EPI_ISL_60 | ···················· | ···················· | ····················· | ···················· | ···················· | ···················· |
| N | 4556 | hCoV-19/Switzerland/GE-ETHZ-321543/2020\|EPI_ISL_60 | ···················· | ···················· | ····················· | ···················· | ···················· | ···················· |
| N | 4557 | hCoV-19/Switzerland/VD-ETHZ-321544/2020\|EPI_ISL_60 | ···················· | ···················· | ····················· | ···················· | ···················· | ···················· |
| N | 4558 | hCoV-19/Switzerland/VD-ETHZ-321545/2020\|EPI_ISL_60 | ···················· | ···················· | ····················· | ···················· | ···················· | ···················· |
| N | 4559 | hCoV-19/Switzerland/VD-ETHZ-321546/2020\|EPI_ISL_60 | ···················· | ···················· | ····················· | ···················· | ···················· | ···················· |
| N | 4560 | hCoV-19/Switzerland/BE-ETHZ-321547/2020\|EPI_ISL_60 | ···················· | ···················· | ····················· | ···················· | ···················· | ···················· |
| N | 4561 | hCoV-19/Switzerland/VD-ETHZ-321549/2020\|EPI_ISL_60 | ···················· | ···················· | ····················· | ···················· | ···················· | ···················· |
| N | 4562 | hCoV-19/USA/AZ-QDX-1549/2020\|EPI_ISL_603759\|2020-0 | ···················· | ···················· | ····················· | ···················· | ···················· | ···················· |
| N | 4563 | hCoV-19/USA/AZ-QDX-1556/2020\|EPI_ISL_603760\|2020-0 | ···················· | ···················· | ····················· | ···················· | ···················· | ···················· |
| N | 4564 | hCoV-19/USA/CA-QDX-1491/2020\|EPI_ISL_603761\|2020-0 | ···················· | ···················· | ····················· | ···················· | ···················· | ···················· |
| N | 4565 | hCoV-19/USA/CA-QDX-1492/2020\|EPI_ISL_603762\|2020-0 | ···················· | ···················· | ····················· | ···················· | ···················· | ···················· |
| N | 4566 | hCoV-19/USA/CA-QDX-1493/2020\|EPI_ISL_603763\|2020-0 | ···················· | ···················· | ····················· | ···················· | ···················· | ···················· |
| N | 4567 | hCoV-19/USA/CA-QDX-1510/2020\|EPI_ISL_603764\|2020-0 | ···················· | ···················· | ····················· | ···················· | ···················· | ···················· |
| N | 4568 | hCoV-19/USA/CA-QDX-1511/2020\|EPI_ISL_603765\|2020-0 | ···················· | ···················· | ····················· | ···················· | ···················· | ···················· |
| N | 4569 | hCoV-19/USA/CA-QDX-1512/2020\|EPI_ISL_603766\|2020-0 | ···················· | ···················· | ····················· | ···················· | ···················· | ···················· |
| N | 4570 | hCoV-19/USA/CA-QDX-1513/2020\|EPI_ISL_603767\|2020-0 | ···················· | ···················· | ····················· | ···················· | ···················· | ···················· |
| N | 4571 | hCoV-19/USA/CA-QDX-1515/2020\|EPI_ISL_603768\|2020-0 | ···················· | ···················· | ····················· | ···················· | ···················· | ···················· |
| N | 4572 | hCoV-19/USA/CA-QDX-1516/2020\|EPI_ISL_603769\|2020-0 | ···················· | ···················· | ····················· | ···················· | ···················· | ···················· |
| N | 4573 | hCoV-19/USA/CA-QDX-1542/2020\|EPI_ISL_603790\|2020-0 | ···················· | ···················· | ····················· | ···················· | ···················· | ···················· |
| N | 4574 | hCoV-19/USA/CA-QDX-1543/2020\|EPI_ISL_603791\|2020-0 | ···················· | ···················· | ····················· | ···················· | ···················· | ···················· |
| N | 4575 | hCoV-19/USA/CA-QDX-1544/2020\|EPI_ISL_603792\|2020-0 | ···················· | ···················· | ····················· | ···················· | ···················· | ···················· |
| N | 4576 | hCoV-19/USA/CA-QDX-1546/2020\|EPI_ISL_603793\|2020-0 | ···················· | ····A··············· | ····················· | ···················· | ···················· | ···················· |
| N | 4577 | hCoV-19/USA/CA-QDX-1547/2020\|EPI_ISL_603794\|2020-0 | ···················· | ···················· | ····················· | ···················· | ···················· | ···················· |
| N | 4578 | hCoV-19/USA/CA-QDX-1548/2020\|EPI_ISL_603795\|2020-0 | ···················· | ···················· | ····················· | ···················· | ···················· | ···················· |
| N | 4579 | hCoV-19/USA/CA-QDX-1550/2020\|EPI_ISL_603796\|2020-0 | ···················· | ···················· | ····················· | ···················· | ···················· | ···················· |
| N | 4580 | hCoV-19/USA/CA-QDX-1551/2020\|EPI_ISL_603797\|2020-0 | ···················· | ···················· | ····················· | ···················· | ···················· | ···················· |
| N | 4581 | hCoV-19/USA/CA-QDX-1552/2020\|EPI_ISL_603798\|2020-0 | ···················· | ···················· | ····················· | ···················· | ···················· | ···················· |
| N | 4582 | hCoV-19/USA/CA-QDX-1553/2020\|EPI_ISL_603799\|2020-0 | ···················· | ···················· | ····················· | ···················· | ···················· | ···················· |
| N | 4583 | hCoV-19/USA/CA-QDX-1554/2020\|EPI_ISL_603800\|2020-0 | ···················· | ···················· | ····················· | ···················· | ···················· | ···················· |
| N | 4584 | hCoV-19/USA/CA-QDX-1555/2020\|EPI_ISL_603801\|2020-0 | ···················· | ···················· | ····················· | ···················· | ···················· | ···················· |
| N | 4585 | hCoV-19/USA/CA-QDX-1557/2020\|EPI_ISL_603802\|2020-0 | ···················· | ···················· | ····················· | ···················· | ···················· | ···················· |

| | | | | | | | | |
|---|---|---|---|---|---|---|---|---|
| N | 4586 | hCoV-19/USA/CA-QDX-1558/2020\|EPI_ISL_603803\|2020-0 | ···················· | ···················· | ····················· | ···················· | ···················· | ···················· |
| N | 4587 | hCoV-19/USA/CO-QDX-1529/2020\|EPI_ISL_603804\|2020-0 | ···················· | ···················· | ····················· | ···················· | ···················· | ···················· |
| N | 4588 | hCoV-19/USA/CT-QDX-1449/2020\|EPI_ISL_603805\|2020-0 | ···················· | ···················· | ····················· | ···················· | ···················· | ···················· |
| N | 4589 | hCoV-19/USA/CT-QDX-1450/2020\|EPI_ISL_603806\|2020-0 | ···················· | ···················· | ····················· | ···················· | ···················· | ···················· |
| N | 4590 | hCoV-19/USA/CT-QDX-1451/2020\|EPI_ISL_603807\|2020-0 | ···················· | ···················· | ····················· | ···················· | ···················· | ···················· |
| N | 4591 | hCoV-19/USA/CT-QDX-1454/2020\|EPI_ISL_603808\|2020-0 | ···················· | ···················· | ····················· | ···················· | ···················· | ···················· |
| N | 4592 | hCoV-19/USA/CT-QDX-1488/2020\|EPI_ISL_603809\|2020-0 | ···················· | ···················· | ····················· | ···················· | ···················· | ···················· |
| N | 4593 | hCoV-19/USA/CT-QDX-1489/2020\|EPI_ISL_603810\|2020-0 | ···················· | ···················· | ····················· | ···················· | ···················· | ···················· |
| N | 4594 | hCoV-19/USA/CT-QDX-1504/2020\|EPI_ISL_603811\|2020-0 | ···················· | ···················· | ····················· | ···················· | ···················· | ···················· |
| N | 4595 | hCoV-19/USA/CT-QDX-1507/2020\|EPI_ISL_603812\|2020-0 | ···················· | ···················· | ····················· | ···················· | ···················· | ···················· |
| N | 4596 | hCoV-19/USA/CT-QDX-1530/2020\|EPI_ISL_603813\|2020-0 | ···················· | ···················· | ····················· | ···················· | ···················· | ···················· |
| N | 4597 | hCoV-19/USA/ID-QDX-1531/2020\|EPI_ISL_603814\|2020-0 | ···················· | ···················· | ····················· | ···················· | ···················· | ···················· |
| N | 4598 | hCoV-19/USA/ID-QDX-1532/2020\|EPI_ISL_603815\|2020-0 | ···················· | ···················· | ····················· | ···················· | ···················· | ···················· |
| N | 4599 | hCoV-19/USA/ID-QDX-1533/2020\|EPI_ISL_603816\|2020-0 | ···················· | ···················· | ····················· | ···················· | ···················· | ···················· |
| N | 4600 | hCoV-19/USA/MA-QDX-1446/2020\|EPI_ISL_603817\|2020-0 | ···················· | ···················· | ····················· | ···················· | ···················· | ···················· |
| N | 4601 | hCoV-19/USA/MA-QDX-1447/2020\|EPI_ISL_603818\|2020-0 | ···················· | ···················· | ····················· | ···················· | ···················· | ···················· |
| N | 4602 | hCoV-19/USA/MA-QDX-1448/2020\|EPI_ISL_603819\|2020-0 | ···················· | ···················· | ····················· | ···················· | ···················· | ···················· |
| N | 4603 | hCoV-19/USA/MA-QDX-1452/2020\|EPI_ISL_603820\|2020-0 | ···················· | ···················· | ····················· | ···················· | ···················· | ···················· |
| N | 4604 | hCoV-19/USA/MA-QDX-1453/2020\|EPI_ISL_603821\|2020-0 | ···T················ | ···················· | ····················· | ···················· | ···················· | ···················· |
| N | 4605 | hCoV-19/USA/MA-QDX-1455/2020\|EPI_ISL_603822\|2020-0 | ···················· | ···················· | ····················· | ···················· | ···················· | ···················· |
| N | 4606 | hCoV-19/USA/MA-QDX-1456/2020\|EPI_ISL_603823\|2020-0 | ···················· | ···················· | ····················· | ···················· | ···················· | ···················· |
| N | 4607 | hCoV-19/USA/MA-QDX-1457/2020\|EPI_ISL_603824\|2020-0 | ···················· | ···················· | ····················· | ···················· | ···················· | ···················· |
| N | 4608 | hCoV-19/USA/UT-QDX-1475/2020\|EPI_ISL_603860\|2020-0 | ···················· | ···················· | ····················· | ···················· | ···················· | ···················· |
| N | 4609 | hCoV-19/England/QEUH-A5E160/2020\|EPI_ISL_597420\|20 | ···················· | ···················· | ····················· | ···················· | ···················· | ···················· |
| N | 4610 | hCoV-19/England/QEUH-A5E0BE/2020\|EPI_ISL_597421\|20 | ···················· | ···················· | ····················· | ···················· | ···················· | ···················· |
| N | 4611 | hCoV-19/England/QEUH-A5DD5E/2020\|EPI_ISL_597434\|20 | ···················· | ···················· | ····················· | ···················· | ···················· | ···················· |
| N | 4612 | hCoV-19/England/CAMC-A57A99/2020\|EPI_ISL_597435\|20 | ···················· | ···················· | ····················· | ···················· | ···················· | ···················· |
| N | 4613 | hCoV-19/England/QEUH-A5E555/2020\|EPI_ISL_597436\|20 | ···················· | ···················· | ····················· | ···················· | ···················· | ···················· |
| N | 4614 | hCoV-19/England/CAMC-A59C37/2020\|EPI_ISL_597437\|20 | ···················· | ···················· | ····················· | ···················· | ···················· | ···················· |
| N | 4615 | hCoV-19/England/CAMC-A578F9/2020\|EPI_ISL_597438\|20 | ···················· | ···················· | ····················· | ···················· | ···················· | ···················· |
| N | 4616 | hCoV-19/England/QEUH-A5E036/2020\|EPI_ISL_597439\|20 | ···················· | ···················· | ····················· | ···················· | ···················· | ···················· |
| N | 4617 | hCoV-19/England/QEUH-A5E1CA/2020\|EPI_ISL_597440\|20 | ···················· | ···················· | ····················· | ···················· | ···················· | ···················· |
| N | 4618 | hCoV-19/England/CAMC-A57149/2020\|EPI_ISL_597441\|20 | ···················· | ···················· | ····················· | ···················· | ···················· | ···················· |
| N | 4619 | hCoV-19/England/QEUH-A5E79B/2020\|EPI_ISL_597442\|20 | ···················· | ···················· | ····················· | ···················· | ···················· | ···················· |
| N | 4620 | hCoV-19/England/QEUH-A5E546/2020\|EPI_ISL_597443\|20 | ···················· | ···················· | ····················· | ···················· | ···················· | ···················· |
| N | 4621 | hCoV-19/England/QEUH-A5E3C4/2020\|EPI_ISL_597444\|20 | ···················· | ···················· | ····················· | ···················· | ···················· | ···················· |
| N | 4622 | hCoV-19/England/CAMC-A57B4B/2020\|EPI_ISL_597445\|20 | ···················· | ···················· | ····················· | ···················· | ···················· | ···················· |
| N | 4623 | hCoV-19/England/CAMC-A5712B/2020\|EPI_ISL_597446\|20 | ···················· | ···················· | ····················· | ···················· | ···················· | ···················· |
| N | 4624 | hCoV-19/England/QEUH-A5E221/2020\|EPI_ISL_597447\|20 | ···················· | ···················· | ····················· | ···················· | ···················· | ···················· |
| N | 4625 | hCoV-19/England/QEUH-A5E591/2020\|EPI_ISL_597448\|20 | ···················· | ···················· | ····················· | ···················· | ···················· | ···················· |
| N | 4626 | hCoV-19/England/CAMC-A579D8/2020\|EPI_ISL_597450\|20 | ···················· | ···················· | ····················· | ···················· | ···················· | ···················· |
| N | 4627 | hCoV-19/England/CAMC-A5747D/2020\|EPI_ISL_597451\|20 | ···················· | ···················· | ····················· | ···················· | ···················· | ···················· |
| N | 4628 | hCoV-19/England/QEUH-A5E810/2020\|EPI_ISL_597452\|20 | ···················· | ···················· | ····················· | ···················· | ···················· | ···················· |
| N | 4629 | hCoV-19/England/CAMC-A57334/2020\|EPI_ISL_597453\|20 | ···················· | ···················· | ····················· | ···················· | ···················· | ···················· |
| N | 4630 | hCoV-19/England/QEUH-A5E2F4/2020\|EPI_ISL_597454\|20 | ···················· | ·A·················· | ····················· | ···················· | ···················· | ··A················· |
| N | 4631 | hCoV-19/England/QEUH-A5DDE5/2020\|EPI_ISL_597455\|20 | ···················· | ···················· | ····················· | ···················· | ···················· | ···················· |
| N | 4632 | hCoV-19/England/QEUH-A5E78C/2020\|EPI_ISL_597456\|20 | ···················· | ···················· | ····················· | ···················· | ···················· | ···················· |
| N | 4633 | hCoV-19/England/CAMC-A573CB/2020\|EPI_ISL_597458\|20 | ···················· | ···················· | ····················· | ···················· | ···················· | ···················· |
| N | 4634 | hCoV-19/England/CAMC-A57185/2020\|EPI_ISL_597460\|20 | ···················· | ···················· | ····················· | ···················· | ···················· | ···················· |
| N | 4635 | hCoV-19/England/CAMC-A59C19/2020\|EPI_ISL_597464\|20 | ···················· | ···················· | ····················· | ···················· | ···················· | ···················· |
| N | 4636 | hCoV-19/England/CAMC-A57B0F/2020\|EPI_ISL_597466\|20 | ···················· | ···················· | ····················· | ···················· | ···················· | ···················· |
| N | 4637 | hCoV-19/England/CAMC-A57774/2020\|EPI_ISL_597467\|20 | ···················· | ···················· | ····················· | ···················· | ···················· | ···················· |
| N | 4638 | hCoV-19/England/CAMC-A57228/2020\|EPI_ISL_597468\|20 | ···················· | ···················· | ····················· | ···················· | ···················· | ···················· |
| N | 4639 | hCoV-19/England/CAMC-A57844/2020\|EPI_ISL_597469\|20 | ···················· | ···················· | ····················· | ···················· | ···················· | ···················· |
| N | 4640 | hCoV-19/England/CAMC-A571D0/2020\|EPI_ISL_597470\|20 | ···················· | ···················· | ····················· | ···················· | ···················· | ···················· |
| N | 4641 | hCoV-19/England/QEUH-A5DFB2/2020\|EPI_ISL_597471\|20 | ···················· | ···················· | ····················· | ···················· | ···················· | ···················· |
| N | 4642 | hCoV-19/England/CAMC-A57862/2020\|EPI_ISL_597472\|20 | ···················· | ···················· | ····················· | ···················· | ···················· | ···················· |
| N | 4643 | hCoV-19/England/CAMC-A571A3/2020\|EPI_ISL_597473\|20 | ···················· | ···················· | ····················· | ···················· | ···················· | ···················· |
| N | 4644 | hCoV-19/England/QEUH-A5E93B/2020\|EPI_ISL_597474\|20 | ···················· | ···················· | ····················· | ···················· | ···················· | ···················· |
| N | 4645 | hCoV-19/England/QEUH-A5E42B/2020\|EPI_ISL_597475\|20 | ···················· | ···················· | ····················· | ···················· | ···················· | ···················· |
| N | 4646 | hCoV-19/England/CAMC-A577FC/2020\|EPI_ISL_597476\|20 | ···················· | ···················· | ····················· | ···················· | ···················· | ···················· |
| N | 4647 | hCoV-19/England/CAMC-A57413/2020\|EPI_ISL_597477\|20 | ···················· | ···················· | ····················· | ···················· | ···················· | ···················· |
| N | 4648 | hCoV-19/England/CAMC-A57932/2020\|EPI_ISL_597478\|20 | ···················· | ···················· | ····················· | ···················· | ···················· | ···················· |
| N | 4649 | hCoV-19/England/CAMC-A579E7/2020\|EPI_ISL_597479\|20 | ···················· | ···················· | ····················· | ···················· | ···················· | ···················· |
| N | 4650 | hCoV-19/England/CAMC-A57765/2020\|EPI_ISL_597480\|20 | ···················· | ···················· | ····················· | ···················· | ···················· | ···················· |
| N | 4651 | hCoV-19/England/QEUH-A5E6AD/2020\|EPI_ISL_597481\|20 | ···················· | ···················· | ····················· | ···················· | ···················· | ···················· |
| N | 4652 | hCoV-19/England/QEUH-A5E6CB/2020\|EPI_ISL_597484\|20 | ···················· | ···················· | ····················· | ···················· | ···················· | ···················· |
| N | 4653 | hCoV-19/England/QEUH-A5E26D/2020\|EPI_ISL_597485\|20 | ···················· | ···················· | ····················· | ···················· | ···················· | ···················· |
| N | 4654 | hCoV-19/England/CAMC-A571C1/2020\|EPI_ISL_597487\|20 | ···················· | ···················· | ····················· | ···················· | ···················· | ···················· |
| N | 4655 | hCoV-19/England/CAMC-A578CC/2020\|EPI_ISL_597488\|20 | ···················· | ···················· | ····················· | ···················· | ···················· | ···················· |
| N | 4656 | hCoV-19/England/CAMC-A57158/2020\|EPI_ISL_597489\|20 | ···················· | ···················· | ····················· | ···················· | ···················· | ···················· |
| N | 4657 | hCoV-19/England/CAMC-A578DB/2020\|EPI_ISL_597490\|20 | ···················· | ···················· | ····················· | ···················· | ···················· | ···················· |
| N | 4658 | hCoV-19/England/QEUH-A5E968/2020\|EPI_ISL_597491\|20 | ···················· | ···················· | ····················· | ···················· | ···················· | ···················· |
| N | 4659 | hCoV-19/England/QEUH-A5E3F1/2020\|EPI_ISL_597492\|20 | ···················· | ···················· | ····················· | ···················· | ···················· | ···················· |
| N | 4660 | hCoV-19/England/QEUH-A5DED3/2020\|EPI_ISL_597493\|20 | ···················· | ···················· | ····················· | ···················· | ···················· | ···················· |
| N | 4661 | hCoV-19/England/CAMC-A576FF/2020\|EPI_ISL_597494\|20 | ···················· | ···················· | ····················· | ···················· | ···················· | ···················· |
| N | 4662 | hCoV-19/England/QEUH-A5E34C/2020\|EPI_ISL_597496\|20 | ···················· | ···················· | ····················· | ···················· | ···················· | ···················· |
| N | 4663 | hCoV-19/England/CAMC-A577CF/2020\|EPI_ISL_597497\|20 | ···················· | ···················· | ····················· | ···················· | ···················· | ···················· |
| N | 4664 | hCoV-19/England/QEUH-A5E5CE/2020\|EPI_ISL_597498\|20 | ···················· | ···················· | ····················· | ···················· | ···················· | ···················· |
| N | 4665 | hCoV-19/England/QEUH-A5E564/2020\|EPI_ISL_597499\|20 | ···················· | ···················· | ····················· | ···················· | ···················· | ···················· |

| | | | | | | | | |
|---|---|---|---|---|---|---|---|---|
| N | 4666 | hCoV-19/England/MILK-A45A25/2020\|EPI_ISL_598700\|20 | ···················· | ···················· | ···················· | ···················· | ···················· | ···················· |
| N | 4667 | hCoV-19/England/MILK-A45C10/2020\|EPI_ISL_598702\|20 | ···················· | ···················· | ···················· | ···················· | ···················· | ···················· |
| N | 4668 | hCoV-19/England/MILK-A46365/2020\|EPI_ISL_598704\|20 | ···················· | ···················· | ···················· | ···················· | ···················· | ···················· |
| N | 4669 | hCoV-19/England/MILK-A4673C/2020\|EPI_ISL_598705\|20 | ···················· | ···················· | ···················· | ···················· | ···················· | ···················· |
| N | 4670 | hCoV-19/England/MILK-A4509C/2020\|EPI_ISL_598706\|20 | ···················· | ···················· | ···················· | ···················· | ···················· | ···················· |
| N | 4671 | hCoV-19/England/MILK-A45B8C/2020\|EPI_ISL_598707\|20 | ···················· | ···················· | ···················· | ···················· | ···················· | ···················· |
| N | 4672 | hCoV-19/England/MILK-A458B2/2020\|EPI_ISL_598708\|20 | ···················· | ···················· | ···················· | ···················· | ···················· | ···················· |
| N | 4673 | hCoV-19/England/MILK-A46778/2020\|EPI_ISL_598709\|20 | ···················· | ···················· | ···················· | ···················· | ···················· | ···················· |
| N | 4674 | hCoV-19/England/MILK-A46787/2020\|EPI_ISL_598710\|20 | ···················· | ···················· | ···················· | ···················· | ·····T·············· | ···················· |
| N | 4675 | hCoV-19/England/MILK-A44E1B/2020\|EPI_ISL_598711\|20 | ···················· | ···················· | ···················· | ···················· | ···················· | ···················· |
| N | 4676 | hCoV-19/England/MILK-A46444/2020\|EPI_ISL_598712\|20 | ···················· | ···················· | ···················· | ···················· | ···················· | ···················· |
| N | 4677 | hCoV-19/England/MILK-A45A43/2020\|EPI_ISL_598713\|20 | ···················· | ·A·················· | ···················· | ···················· | ···················· | ···················· |
| N | 4678 | hCoV-19/England/MILK-A45D77/2020\|EPI_ISL_598714\|20 | ···················· | ···················· | ···················· | ···················· | ···················· | ···················· |
| N | 4679 | hCoV-19/England/MILK-A45A52/2020\|EPI_ISL_598715\|20 | ···················· | ···················· | ···················· | ···················· | ···················· | ···················· |
| N | 4680 | hCoV-19/England/MILK-A45A34/2020\|EPI_ISL_598716\|20 | ···················· | ···················· | ···················· | ···················· | ···················· | ···················· |
| N | 4681 | hCoV-19/England/MILK-A45CD4/2020\|EPI_ISL_598717\|20 | ···················· | ···················· | ···················· | ···················· | ···················· | ···················· |
| N | 4682 | hCoV-19/England/MILK-A45928/2020\|EPI_ISL_598718\|20 | ···················· | ···················· | ···················· | ···················· | ···················· | ···················· |
| N | 4683 | hCoV-19/England/MILK-A45F44/2020\|EPI_ISL_598719\|20 | ···················· | ···················· | ···················· | ···················· | ···················· | ···················· |
| N | 4684 | hCoV-19/England/MILK-A45B04/2020\|EPI_ISL_598720\|20 | ···················· | ···················· | ···················· | ···················· | ···················· | ···················· |
| N | 4685 | hCoV-19/England/MILK-A44FDC/2020\|EPI_ISL_598750\|20 | ···················· | ···················· | ···················· | ···················· | ···················· | ···················· |
| N | 4686 | hCoV-19/England/MILK-A45ABC/2020\|EPI_ISL_598752\|20 | ···················· | ···················· | ···················· | ···················· | ···················· | ···················· |
| N | 4687 | hCoV-19/England/MILK-A46857/2020\|EPI_ISL_598767\|20 | ···················· | ···················· | ···················· | ···················· | ···················· | ···················· |
| N | 4688 | hCoV-19/England/MILK-A4663F/2020\|EPI_ISL_598769\|20 | ···················· | ···················· | ···················· | ···················· | ···················· | ···················· |
| N | 4689 | hCoV-19/England/MILK-A457A6/2020\|EPI_ISL_598770\|20 | ···················· | ···················· | ···················· | ···················· | ···················· | ···················· |
| N | 4690 | hCoV-19/England/MILK-A467E1/2020\|EPI_ISL_598771\|20 | ···················· | ···················· | ···················· | ···················· | ···················· | ···················· |
| N | 4691 | hCoV-19/England/MILK-A45E1A/2020\|EPI_ISL_598772\|20 | ···················· | ···················· | ···················· | ···················· | ···················· | ···················· |
| N | 4692 | hCoV-19/England/MILK-A4617A/2020\|EPI_ISL_598773\|20 | ···················· | ···················· | ···················· | ···················· | ···················· | ···················· |
| N | 4693 | hCoV-19/England/MILK-A45E74/2020\|EPI_ISL_598774\|20 | ···················· | ···················· | ···················· | ···················· | ···················· | ···················· |
| N | 4694 | hCoV-19/England/MILK-A45D2C/2020\|EPI_ISL_598775\|20 | ···················· | ·A·················· | ···················· | ···················· | ···················· | ··A················· |
| N | 4695 | hCoV-19/England/MILK-A45A07/2020\|EPI_ISL_598776\|20 | ···················· | ···················· | ···················· | ···················· | ···················· | ···················· |
| N | 4696 | hCoV-19/England/MILK-A45A70/2020\|EPI_ISL_598778\|20 | ···················· | ···················· | ···················· | ···················· | ···················· | ···················· |
| N | 4697 | hCoV-19/England/MILK-A460C8/2020\|EPI_ISL_598779\|20 | ···················· | ···················· | ···················· | ···················· | ···················· | ···················· |
| N | 4698 | hCoV-19/England/MILK-A45DB3/2020\|EPI_ISL_598780\|20 | ···················· | ···················· | ···················· | ···················· | ···················· | ···················· |
| N | 4699 | hCoV-19/England/MILK-A44E84/2020\|EPI_ISL_598781\|20 | ···················· | ···················· | ···················· | ···················· | ···················· | ···················· |
| N | 4700 | hCoV-19/England/MILK-A44D1E/2020\|EPI_ISL_598782\|20 | ···················· | ···················· | ···················· | ···················· | ·····T·············· | ···················· |
| N | 4701 | hCoV-19/England/MILK-A45CA7/2020\|EPI_ISL_598783\|20 | ···················· | ···················· | ···················· | ···················· | ···················· | ···················· |
| N | 4702 | hCoV-19/England/MILK-A45ECF/2020\|EPI_ISL_598784\|20 | ···················· | ···················· | ···················· | ···················· | ···················· | ···················· |
| N | 4703 | hCoV-19/England/MILK-A45937/2020\|EPI_ISL_598785\|20 | ···················· | ···················· | ···················· | ···················· | ···················· | ···················· |
| N | 4704 | hCoV-19/England/MILK-A44FBE/2020\|EPI_ISL_598786\|20 | ···················· | ···················· | ···················· | ···················· | ···················· | ···················· |
| N | 4705 | hCoV-19/England/MILK-A457D3/2020\|EPI_ISL_598787\|20 | ···················· | ·A·················· | ···················· | ···················· | ···················· | ···················· |
| N | 4706 | hCoV-19/England/MILK-A4681B/2020\|EPI_ISL_598788\|20 | ···················· | ···················· | ···················· | ···················· | ···················· | ···················· |
| N | 4707 | hCoV-19/England/MILK-A44E39/2020\|EPI_ISL_598789\|20 | ···················· | ···················· | ···················· | ···················· | ···················· | ···················· |
| N | 4708 | hCoV-19/England/MILK-A45D0E/2020\|EPI_ISL_598790\|20 | ···················· | ·A·················· | ···················· | ···················· | ···················· | ··A················· |
| N | 4709 | hCoV-19/England/MILK-A45F80/2020\|EPI_ISL_598791\|20 | ···················· | ···················· | ···················· | ···················· | ···················· | ···················· |
| N | 4710 | hCoV-19/England/MILK-A466D5/2020\|EPI_ISL_598792\|20 | ···················· | ···················· | ···················· | ···················· | ···················· | ···················· |
| N | 4711 | hCoV-19/England/MILK-A45EED/2020\|EPI_ISL_598794\|20 | ···················· | ···················· | ···················· | ···················· | ···················· | ···················· |
| N | 4712 | hCoV-19/England/MILK-A45919/2020\|EPI_ISL_598795\|20 | ···················· | ···················· | ···················· | ···················· | ···················· | ···················· |
| N | 4713 | hCoV-19/England/MILK-A45FEA/2020\|EPI_ISL_598796\|20 | ···················· | ·R·················· | ···················· | ···················· | ···················· | ··R················· |
| N | 4714 | hCoV-19/England/MILK-A45B22/2020\|EPI_ISL_598797\|20 | ···················· | ···················· | ···················· | ···················· | ···················· | ···················· |
| N | 4715 | hCoV-19/England/MILK-A45946/2020\|EPI_ISL_598798\|20 | ···················· | ···················· | ···················· | ···················· | ···················· | ···················· |
| N | 4716 | hCoV-19/England/MILK-A44B8D/2020\|EPI_ISL_598799\|20 | ···················· | ···················· | ···················· | ···················· | ·····T·············· | ···················· |
| N | 4717 | hCoV-19/Scotland/QEUH-9DCF07/2020\|EPI_ISL_601111\|2 | ···················· | ···················· | ···················· | ···················· | ···················· | ···················· |
| N | 4718 | hCoV-19/Scotland/QEUH-9DC15C/2020\|EPI_ISL_601112\|2 | ···················· | ···················· | ···················· | ···················· | ···················· | ···················· |
| N | 4719 | hCoV-19/Scotland/QEUH-9DAC6C/2020\|EPI_ISL_601113\|2 | ···················· | ···················· | ···················· | ···················· | ···················· | ···················· |
| N | 4720 | hCoV-19/Scotland/QEUH-9DADA5/2020\|EPI_ISL_601114\|2 | ···················· | ···················· | ···················· | ···················· | ···················· | ···················· |
| N | 4721 | hCoV-19/Scotland/QEUH-9DC356/2020\|EPI_ISL_601115\|2 | ···················· | ···················· | ···················· | ···················· | ···················· | ···················· |
| N | 4722 | hCoV-19/Scotland/QEUH-9DAB32/2020\|EPI_ISL_601116\|2 | ···················· | ···················· | ···················· | ···················· | ···················· | ···················· |
| N | 4723 | hCoV-19/Scotland/QEUH-9DCAD9/2020\|EPI_ISL_601117\|2 | ···················· | ···················· | ···················· | ···················· | ···················· | ···················· |
| N | 4724 | hCoV-19/England/QEUH-9DA956/2020\|EPI_ISL_601118\|20 | ···················· | ···················· | ···················· | ···················· | ···················· | ···················· |
| N | 4725 | hCoV-19/Scotland/QEUH-9DAD96/2020\|EPI_ISL_601119\|2 | ···················· | ···················· | ···················· | ···················· | ···················· | ···················· |
| N | 4726 | hCoV-19/Scotland/QEUH-9DAEEE/2020\|EPI_ISL_601120\|2 | ···················· | ···················· | ···················· | ···················· | ···················· | ···················· |
| N | 4727 | hCoV-19/Scotland/QEUH-9DC9FA/2020\|EPI_ISL_601121\|2 | ···················· | ···················· | ···················· | ···················· | ···················· | ···················· |
| N | 4728 | hCoV-19/Scotland/QEUH-9DCE73/2020\|EPI_ISL_601122\|2 | ···················· | ···················· | ···················· | ···················· | ···················· | ···················· |
| N | 4729 | hCoV-19/Scotland/QEUH-9DAB9C/2020\|EPI_ISL_601123\|2 | ···················· | ···················· | ···················· | ···················· | ···················· | ···················· |
| N | 4730 | hCoV-19/Scotland/QEUH-9DACC6/2020\|EPI_ISL_601124\|2 | ···················· | ···················· | ···················· | ···················· | ···················· | ···················· |
| N | 4731 | hCoV-19/Scotland/QEUH-9DC4F9/2020\|EPI_ISL_601125\|2 | ···················· | ···················· | ···················· | ···················· | ···················· | ···················· |
| N | 4732 | hCoV-19/Scotland/QEUH-9DC16B/2020\|EPI_ISL_601126\|2 | ···················· | ···················· | ···················· | ···················· | ···················· | ···················· |
| N | 4733 | hCoV-19/Scotland/QEUH-9DCB12/2020\|EPI_ISL_601127\|2 | ···················· | ···················· | ···················· | ···················· | ···················· | ···················· |
| N | 4734 | hCoV-19/Scotland/QEUH-9DB005/2020\|EPI_ISL_601128\|2 | ···················· | ···················· | ···················· | ···················· | ···················· | ···················· |
| N | 4735 | hCoV-19/Scotland/QEUH-9DC9DC/2020\|EPI_ISL_601129\|2 | ···················· | ···················· | ···················· | ···················· | ···················· | ···················· |
| N | 4736 | hCoV-19/Scotland/QEUH-9DABAB/2020\|EPI_ISL_601170\|2 | ···················· | ···················· | ···················· | ···················· | ···················· | ···················· |
| N | 4737 | hCoV-19/Scotland/QEUH-9DABC9/2020\|EPI_ISL_601171\|2 | ···················· | ···················· | ···················· | ···················· | ···················· | ···················· |
| N | 4738 | hCoV-19/Scotland/QEUH-9DC9EB/2020\|EPI_ISL_601172\|2 | ···················· | ···················· | ···················· | ···················· | ···················· | ···················· |
| N | 4739 | hCoV-19/Scotland/QEUH-9DCECE/2020\|EPI_ISL_601173\|2 | ···················· | ···················· | ···················· | ···················· | ···················· | ···················· |
| N | 4740 | hCoV-19/Scotland/QEUH-9DC4DB/2020\|EPI_ISL_601174\|2 | ···················· | ···················· | ···················· | ···················· | ·····T·············· | ···················· |
| N | 4741 | hCoV-19/Scotland/QEUH-9DC462/2020\|EPI_ISL_601175\|2 | ···················· | ···················· | ···················· | ···················· | ···T················ | ···················· |
| N | 4742 | hCoV-19/Scotland/QEUH-9DAF81/2020\|EPI_ISL_601176\|2 | ···················· | ···················· | ···················· | ···················· | ·····T·············· | ···················· |
| N | 4743 | hCoV-19/Scotland/QEUH-9DC82A/2020\|EPI_ISL_601177\|2 | ···················· | ···················· | ···················· | ···················· | ···················· | ···················· |
| N | 4744 | hCoV-19/Scotland/QEUH-9DAD4B/2020\|EPI_ISL_601178\|2 | ···················· | ···················· | ···················· | ···················· | ···················· | ···················· |
| N | 4745 | hCoV-19/Scotland/QEUH-9DAD5A/2020\|EPI_ISL_601179\|2 | ···················· | ···················· | ···················· | ···················· | ···················· | ···················· |

N 4746 hCoV-19/Scotland/QEUH-9DAB23/2020|EPI_ISL_601180|2 ···················· ···················· ····················· ···················· ···················· ····················
N 4747 hCoV-19/Scotland/QEUH-9DC417/2020|EPI_ISL_601181|2 ···················· ···················· ····················· ···················· ·····T·············· ····················
N 4748 hCoV-19/Scotland/QEUH-9DC435/2020|EPI_ISL_601182|2 ···················· ···················· ····················· ···················· ···················· ····················
N 4749 hCoV-19/Scotland/QEUH-9DCB03/2020|EPI_ISL_601183|2 ···················· ···················· ····················· ···················· ···················· ····················
N 4750 hCoV-19/Scotland/QEUH-9DC347/2020|EPI_ISL_601184|2 ···················· ···················· ····················· ···················· ···················· ····················
N 4751 hCoV-19/Scotland/QEUH-9DC453/2020|EPI_ISL_601185|2 ···················· ···················· ····················· ···················· ···················· ····················
N 4752 hCoV-19/Scotland/QEUH-9DB08D/2020|EPI_ISL_601186|2 ···················· ···················· ····················· ···················· ···················· ····················
N 4753 hCoV-19/Scotland/QEUH-9DE891/2020|EPI_ISL_601187|2 ···················· ···················· ····················· ···················· ···················· ····················
N 4754 hCoV-19/Scotland/QEUH-9DE70D/2020|EPI_ISL_601188|2 ···················· ···················· ····················· ···················· ···················· ····················
N 4755 hCoV-19/Scotland/QEUH-9DE7A3/2020|EPI_ISL_601189|2 ···················· ···················· ····················· ···················· ···················· ····················
N 4756 hCoV-19/Scotland/QEUH-9DEAC8/2020|EPI_ISL_601190|2 ···················· ···················· ····················· ···················· ···················· ····················
N 4757 hCoV-19/Scotland/QEUH-9DEA22/2020|EPI_ISL_601191|2 ···················· ···················· ····················· ···················· ···················· ····················
N 4758 hCoV-19/Scotland/QEUH-9DE679/2020|EPI_ISL_601193|2 ···················· ···················· ····················· ···················· ···················· ····················
N 4759 hCoV-19/Scotland/QEUH-9DE794/2020|EPI_ISL_601194|2 ···················· ···················· ····················· ···················· ···················· ····················
N 4760 hCoV-19/Scotland/QEUH-9DE8BF/2020|EPI_ISL_601195|2 ···················· ···················· ····················· ···················· ···················· ····················
N 4761 hCoV-19/Scotland/QEUH-9DEDED/2020|EPI_ISL_601196|2 ···················· ···················· ····················· ···················· ···················· ····················
N 4762 hCoV-19/Scotland/QEUH-9DE6B5/2020|EPI_ISL_601197|2 ···················· ···················· ····················· ···················· ···················· ····················
N 4763 hCoV-19/Scotland/QEUH-9DEDCF/2020|EPI_ISL_601198|2 ···················· ···················· ····················· ···················· ···················· ····················
N 4764 hCoV-19/Scotland/QEUH-9DE688/2020|EPI_ISL_601199|2 ···················· ···················· ····················· ···················· ···················· ····················
N 4765 hCoV-19/Russia/MUR-RII-MH2065S/2020|EPI_ISL_602400 ···················· ···················· ····················· ···················· ···················· ····················
N 4766 hCoV-19/Russia/SVE-RII-MH2066S/2020|EPI_ISL_602401 ···················· ···················· ····················· ···················· ···················· ····················
N 4767 hCoV-19/Russia/SVE-RII-MH2069S/2020|EPI_ISL_602402 ···················· ···················· ····················· ···················· ···················· ····················
N 4768 hCoV-19/Russia/SVE-RII-MH2073S/2020|EPI_ISL_602403 ···················· ···················· ····················· ···················· ···················· ····················
N 4769 hCoV-19/Russia/PER-RII-MH2074S/2020|EPI_ISL_602404 ···················· ···················· ····················· ···················· ···················· ····················
N 4770 hCoV-19/Russia/KR-RII-MH2076S/2020|EPI_ISL_602405| ···················· ···················· ····················· ···················· ···················· ····················
N 4771 hCoV-19/Russia/MUR-RII-MH2077S/2020|EPI_ISL_602406 ···················· ···················· ····················· ···················· ···················· ····················
N 4772 hCoV-19/Russia/MUR-RII-MH2078S/2020|EPI_ISL_602407 ···················· ···················· ····················· ···················· ···················· ····················
N 4773 hCoV-19/Russia/MUR-RII-MH2079S/2020|EPI_ISL_602408 ···················· ···················· ····················· ···················· ···················· ····················
N 4774 hCoV-19/Russia/KO-RII-MH2080S/2020|EPI_ISL_602409| ···················· ···················· ····················· ···················· ···················· ····················
N 4775 hCoV-19/Russia/LEN-RII-MH2081S/2020|EPI_ISL_602410 ···················· ···················· ····················· ···················· ···················· ····················
N 4776 hCoV-19/Russia/SPE-RII-MH2083S/2020|EPI_ISL_602411 ···················· ···················· ····················· ···················· ···················· ····················
N 4777 hCoV-19/Russia/CE-RII-MH2523S/2020|EPI_ISL_602412| ···················· ···················· ····················· ···················· ···················· ····················
N 4778 hCoV-19/Russia/CE-RII-MH2636S/2020|EPI_ISL_602413| ···················· ···················· ····················· ···················· ···················· ····················
N 4779 hCoV-19/Russia/CE-RII-MH2649S/2020|EPI_ISL_602414| ···················· ···················· ····················· ···················· ···················· ····················
N 4780 hCoV-19/Russia/CE-RII-MH2709S/2020|EPI_ISL_602415| ···················· ···················· ····················· ···················· ···················· ····················
N 4781 hCoV-19/Russia/CE-RII-MH2728S/2020|EPI_ISL_602416| ···················· ···················· ····················· ···················· ···················· ····················
N 4782 hCoV-19/Russia/KDA-RII-MH2769S/2020|EPI_ISL_602440 ···················· ···················· ····················· ···················· ···················· ····················
N 4783 hCoV-19/Russia/KDA-RII-MH2770S/2020|EPI_ISL_602441 ···················· ···················· ····················· ···················· ···················· ····················
N 4784 hCoV-19/Russia/KDA-RII-MH2772S/2020|EPI_ISL_602442 ···················· ···················· ····················· ···················· ···················· ····················
N 4785 hCoV-19/Russia/KDA-RII-MH2773S/2020|EPI_ISL_602443 ···················· ···················· ····················· ···················· ···················· ····················
N 4786 hCoV-19/Russia/KDA-RII-MH2774S/2020|EPI_ISL_602444 ···················· ···················· ····················· ···················· ···················· ····················
N 4787 hCoV-19/Russia/STA-RII-MH2777S/2020|EPI_ISL_602445 ···················· ···················· ····················· ···················· ···················· ····················
N 4788 hCoV-19/Russia/STA-RII-MH2780S/2020|EPI_ISL_602446 ···················· ···················· ····················· ···················· ···················· ····················
N 4789 hCoV-19/Russia/STA-RII-MH2782S/2020|EPI_ISL_602447 ···················· ···················· ····················· ···················· ···················· ····················
N 4790 hCoV-19/Russia/STA-RII-MH2783S/2020|EPI_ISL_602448 ···················· ···················· ····················· ···················· ···················· ····················
N 4791 hCoV-19/Russia/STA-RII-MH2784S/2020|EPI_ISL_602449 ···················· ···················· ····················· ···················· ···················· ····················
N 4792 hCoV-19/Russia/STA-RII-MH2785S/2020|EPI_ISL_602450 ···················· ···················· ····················· ···················· ···················· ····················
N 4793 hCoV-19/Russia/STA-RII-MH2786S/2020|EPI_ISL_602451 ···················· ···················· ····················· ···················· ···················· ····················
N 4794 hCoV-19/Russia/STA-RII-MH2787S/2020|EPI_ISL_602452 ···················· ···················· ····················· ···················· ···················· ····················
N 4795 hCoV-19/Russia/BRY-RII-MH2790S/2020|EPI_ISL_602453 ···················· ···················· ····················· ···················· ···················· ····················
N 4796 hCoV-19/Russia/VOR-RII-MH2792S/2020|EPI_ISL_602454 ···················· ···················· ····················· ···················· ···················· ····················
N 4797 hCoV-19/Russia/VOR-RII-MH2793S/2020|EPI_ISL_602455 ···················· ···················· ····················· ···················· ···················· ····················
N 4798 hCoV-19/Russia/IVA-RII-MH2794S/2020|EPI_ISL_602456 ···················· ···················· ····················· ···················· ···················· ····················
N 4799 hCoV-19/Russia/KLU-RII-MH2795S/2020|EPI_ISL_602457 ···················· ···················· ····················· ···················· ···················· ····················
N 4800 hCoV-19/Russia/KEM-RII-MH2796S/2020|EPI_ISL_602458 ···················· ···················· ····················· ···················· ···················· ····················
N 4801 hCoV-19/Russia/MOS-RII-MH2797S/2020|EPI_ISL_602459 ···················· ···················· ····················· ···················· ···················· ····················
N 4802 hCoV-19/Switzerland/BE-ETHZ-321492/2020|EPI_ISL_60 ···················· ···················· ····················· ···················· ···················· ····················
N 4803 hCoV-19/Switzerland/BE-ETHZ-321493/2020|EPI_ISL_60 ···················· ···················· ····················· ···················· ···················· ····················
N 4804 hCoV-19/Switzerland/BE-ETHZ-321494/2020|EPI_ISL_60 ···················· ···················· ····················· ···················· ···················· ····················
N 4805 hCoV-19/Switzerland/VD-ETHZ-321495/2020|EPI_ISL_60 ···················· ···················· ····················· ···················· ···················· ····················
N 4806 hCoV-19/Switzerland/SO-ETHZ-321496/2020|EPI_ISL_60 ···················· ···················· ····················· ···················· ···················· ····················
N 4807 hCoV-19/Switzerland/VD-ETHZ-321498/2020|EPI_ISL_60 ···················· ···················· ····················· ···················· ···················· ····················
N 4808 hCoV-19/Switzerland/VD-ETHZ-321499/2020|EPI_ISL_60 ···················· ···················· ····················· ···················· ···················· ····················
N 4809 hCoV-19/Switzerland/VD-ETHZ-321500/2020|EPI_ISL_60 ···················· ···················· ····················· ···················· ···················· ····················
N 4810 hCoV-19/Switzerland/BL-ETHZ-321501/2020|EPI_ISL_60 ···················· ···················· ····················· ···················· ···················· ····················
N 4811 hCoV-19/Switzerland/VD-ETHZ-321502/2020|EPI_ISL_60 ···················· ···················· ····················· ···················· ···················· ····················
N 4812 hCoV-19/Switzerland/VD-ETHZ-321503/2020|EPI_ISL_60 ···················· ···················· ····················· ···················· ···················· ····················
N 4813 hCoV-19/Switzerland/VD-ETHZ-321505/2020|EPI_ISL_60 ···················· ···················· ····················· ···················· ···················· ····················
N 4814 hCoV-19/Switzerland/VD-ETHZ-321506/2020|EPI_ISL_60 ···················· ···················· ····················· ···················· ···················· ····················
N 4815 hCoV-19/Switzerland/SZ-ETHZ-321507/2020|EPI_ISL_60 ···················· ···················· ····················· ···················· ···················· ····················
N 4816 hCoV-19/Switzerland/AG-ETHZ-321508/2020|EPI_ISL_60 ···················· ···················· ····················· ···················· ···················· ····················
N 4817 hCoV-19/Switzerland/ZH-ETHZ-321509/2020|EPI_ISL_60 ···················· ···················· ····················· ···················· ···················· ····················
N 4818 hCoV-19/Switzerland/ZH-ETHZ-321510/2020|EPI_ISL_60 ···················· ···················· ····················· ···················· ···················· ····················
N 4819 hCoV-19/Switzerland/ZH-ETHZ-321511/2020|EPI_ISL_60 ···················· ···················· ····················· ···················· ···················· ····················
N 4820 hCoV-19/Switzerland/ZH-ETHZ-321512/2020|EPI_ISL_60 ···················· ··········Y········· ····················· ···················· ···················· ····················
N 4821 hCoV-19/Switzerland/AG-ETHZ-321513/2020|EPI_ISL_60 ···················· ···················· ····················· ···················· ···················· ····················
N 4822 hCoV-19/Switzerland/BE-ETHZ-321515/2020|EPI_ISL_60 ···················· ···················· ····················· ···················· ···················· ····················
N 4823 hCoV-19/Switzerland/ZG-ETHZ-321516/2020|EPI_ISL_60 ···················· ···················· ····················· ···················· ···················· ····················
N 4824 hCoV-19/Switzerland/UR-ETHZ-321517/2020|EPI_ISL_60 ···················· ···················· ····················· ···················· ··············K····· ····················
N 4825 hCoV-19/Switzerland/ZG-ETHZ-321518/2020|EPI_ISL_60 ···················· ···················· ····················· ···················· ···················· ····················

| | | | | | | | | |
|---|---|---|---|---|---|---|---|---|
| N | 4826 | hCoV-19/Switzerland/UR-ETHZ-321519/2020\|EPI_ISL_60 | ···················· | ···················· | ···················· | ···················· | ···················· | ···················· |
| N | 4827 | hCoV-19/Switzerland/AG-ETHZ-321520/2020\|EPI_ISL_60 | ···················· | ···················· | ···················· | ···················· | ···················· | ···················· |
| N | 4828 | hCoV-19/Switzerland/UR-ETHZ-321521/2020\|EPI_ISL_60 | ···················· | ···················· | ···················· | ···················· | ···················· | ···················· |
| N | 4829 | hCoV-19/Switzerland/BE-ETHZ-321522/2020\|EPI_ISL_60 | ···················· | ···················· | ···················· | ···················· | ···················· | ···················· |
| N | 4830 | hCoV-19/Switzerland/BS-ETHZ-321523/2020\|EPI_ISL_60 | ···················· | ·A·················· | ···················· | ···················· | ···················· | ···················· |
| N | 4831 | hCoV-19/Switzerland/ZH-ETHZ-321524/2020\|EPI_ISL_60 | ···················· | ···················· | ···················· | ···················· | ···················· | ···················· |
| N | 4832 | hCoV-19/Switzerland/SO-ETHZ-321525/2020\|EPI_ISL_60 | ···················· | ···················· | ···················· | ···················· | ···················· | ···················· |
| N | 4833 | hCoV-19/Switzerland/TI-ETHZ-321526/2020\|EPI_ISL_60 | ···················· | ···················· | ···················· | ···················· | ···················· | ···················· |
| N | 4834 | hCoV-19/Switzerland/ZH-ETHZ-321527/2020\|EPI_ISL_60 | ···················· | ···················· | ···················· | ···················· | ···················· | ···················· |
| N | 4835 | hCoV-19/Switzerland/ZH-ETHZ-321528/2020\|EPI_ISL_60 | ···················· | ···················· | ···················· | ···················· | ···················· | ···················· |
| N | 4836 | hCoV-19/Switzerland/SO-ETHZ-321529/2020\|EPI_ISL_60 | ···················· | ···················· | ···················· | ···················· | ······K············· | ···················· |
| N | 4837 | hCoV-19/Switzerland/SO-ETHZ-321530/2020\|EPI_ISL_60 | ···················· | ···················· | ···················· | ···················· | ···················· | ···················· |
| N | 4838 | hCoV-19/Switzerland/GE-ETHZ-321531/2020\|EPI_ISL_60 | ···················· | ···················· | ···K················ | ···················· | ···················· | ···················· |
| N | 4839 | hCoV-19/Switzerland/VD-ETHZ-321532/2020\|EPI_ISL_60 | ···················· | ···················· | ···················· | ···················· | ···················· | ···················· |
| N | 4840 | hCoV-19/Switzerland/VD-ETHZ-321533/2020\|EPI_ISL_60 | ···················· | ···················· | ···················· | ···················· | ···················· | ···················· |
| N | 4841 | hCoV-19/Switzerland/UR-ETHZ-321534/2020\|EPI_ISL_60 | ···················· | ···················· | ···················· | ···················· | ···················· | ···················· |
| N | 4842 | hCoV-19/Switzerland/UR-ETHZ-321535/2020\|EPI_ISL_60 | ···················· | ···················· | ···················· | ···················· | ···················· | ···················· |
| N | 4843 | hCoV-19/Switzerland/UR-ETHZ-321536/2020\|EPI_ISL_60 | ···················· | ···················· | ···················· | ···················· | ···················· | ···················· |
| N | 4844 | hCoV-19/Switzerland/VD-ETHZ-321537/2020\|EPI_ISL_60 | ···················· | ···················· | ···················· | ···················· | ···················· | ···················· |
| N | 4845 | hCoV-19/Switzerland/VD-ETHZ-321538/2020\|EPI_ISL_60 | ···················· | ···················· | ···················· | ···················· | ···················· | ···················· |
| N | 4846 | hCoV-19/USA/CA-QDX-1517/2020\|EPI_ISL_603770\|2020-0 | ···················· | ···················· | ···················· | ···················· | ···················· | ···················· |
| N | 4847 | hCoV-19/USA/CA-QDX-1518/2020\|EPI_ISL_603771\|2020-0 | ···················· | ···················· | ···················· | ···················· | ···················· | ···················· |
| N | 4848 | hCoV-19/USA/CA-QDX-1519/2020\|EPI_ISL_603772\|2020-0 | ···················· | ···················· | ···················· | ···················· | ···················· | ···················· |
| N | 4849 | hCoV-19/USA/CA-QDX-1520/2020\|EPI_ISL_603773\|2020-0 | ···················· | ···················· | ···················· | ···················· | ···················· | ···················· |
| N | 4850 | hCoV-19/USA/CA-QDX-1521/2020\|EPI_ISL_603774\|2020-0 | ···················· | ···················· | ···················· | ···················· | ···················· | ···················· |
| N | 4851 | hCoV-19/USA/CA-QDX-1522/2020\|EPI_ISL_603775\|2020-0 | ···················· | ···················· | ···················· | ···················· | ···················· | ···················· |
| N | 4852 | hCoV-19/USA/CA-QDX-1523/2020\|EPI_ISL_603776\|2020-0 | ···················· | ···················· | ···················· | ···················· | ···················· | ···················· |
| N | 4853 | hCoV-19/USA/CA-QDX-1524/2020\|EPI_ISL_603777\|2020-0 | ···················· | ···················· | ···················· | ···················· | ···················· | ···················· |
| N | 4854 | hCoV-19/USA/CA-QDX-1525/2020\|EPI_ISL_603778\|2020-0 | ···················· | ···················· | ···················· | ···················· | ···················· | ···················· |
| N | 4855 | hCoV-19/USA/CA-QDX-1526/2020\|EPI_ISL_603779\|2020-0 | ···················· | ···················· | ···················· | ···················· | ···················· | ···················· |
| N | 4856 | hCoV-19/USA/CA-QDX-1527/2020\|EPI_ISL_603780\|2020-0 | ···················· | ···················· | ···················· | ···················· | ···················· | ···················· |
| N | 4857 | hCoV-19/USA/CA-QDX-1528/2020\|EPI_ISL_603781\|2020-0 | ···················· | ···················· | ···················· | ···················· | ···················· | ···················· |
| N | 4858 | hCoV-19/USA/CA-QDX-1534/2020\|EPI_ISL_603782\|2020-0 | ···················· | ···················· | ···················· | ···················· | ···················· | ···················· |
| N | 4859 | hCoV-19/USA/CA-QDX-1535/2020\|EPI_ISL_603783\|2020-0 | ···················· | ···················· | ···················· | ···················· | ···················· | ···················· |
| N | 4860 | hCoV-19/USA/CA-QDX-1536/2020\|EPI_ISL_603784\|2020-0 | ···················· | ···················· | ···················· | ···················· | ···················· | ···················· |
| N | 4861 | hCoV-19/USA/CA-QDX-1537/2020\|EPI_ISL_603785\|2020-0 | ···················· | ···················· | ···················· | ···················· | ···················· | ···················· |
| N | 4862 | hCoV-19/USA/CA-QDX-1538/2020\|EPI_ISL_603786\|2020-0 | ···················· | ···················· | ···················· | ···················· | ···················· | ···················· |
| N | 4863 | hCoV-19/USA/CA-QDX-1540/2020\|EPI_ISL_603788\|2020-0 | ···················· | ···················· | ···················· | ···················· | ···················· | ···················· |
| N | 4864 | hCoV-19/USA/CA-QDX-1541/2020\|EPI_ISL_603789\|2020-0 | ···················· | ···················· | ···················· | ···················· | ···················· | ···················· |
| N | 4865 | hCoV-19/England/QEUH-A5E573/2020\|EPI_ISL_597400\|20 | ···················· | ···················· | ···················· | ···················· | ···················· | ···················· |
| N | 4866 | hCoV-19/England/CAMC-A577A1/2020\|EPI_ISL_597401\|20 | ···················· | ···················· | ···················· | ···················· | ···················· | ···················· |
| N | 4867 | hCoV-19/England/CAMC-A57941/2020\|EPI_ISL_597402\|20 | ···················· | ···················· | ···················· | ···················· | ···················· | ···················· |
| N | 4868 | hCoV-19/England/QEUH-A5E6F8/2020\|EPI_ISL_597404\|20 | ···················· | ···················· | ···················· | ···················· | ···················· | ···················· |
| N | 4869 | hCoV-19/England/CAMC-A57246/2020\|EPI_ISL_597405\|20 | ···················· | ···················· | ···················· | ···················· | ······T············· | ···················· |
| N | 4870 | hCoV-19/England/QEUH-A5DE97/2020\|EPI_ISL_597406\|20 | ···················· | ·A·················· | ···················· | ···················· | ···················· | ···A················ |
| N | 4871 | hCoV-19/England/QEUH-A5E115/2020\|EPI_ISL_597407\|20 | ···················· | ···················· | ···················· | ···················· | ···················· | ···················· |
| N | 4872 | hCoV-19/England/QEUH-A5E986/2020\|EPI_ISL_597408\|20 | ···················· | ···················· | ···················· | ···················· | ···················· | ···················· |
| N | 4873 | hCoV-19/England/QEUH-A5DF1C/2020\|EPI_ISL_597409\|20 | ···················· | ···················· | ···················· | ···················· | ···················· | ···················· |
| N | 4874 | hCoV-19/England/QEUH-A5DD4F/2020\|EPI_ISL_597410\|20 | ···················· | ···················· | ···················· | ···················· | ···················· | ···················· |
| N | 4875 | hCoV-19/England/QEUH-A5E0EB/2020\|EPI_ISL_597411\|20 | ···················· | ···················· | ···················· | ···················· | ···················· | ···················· |
| N | 4876 | hCoV-19/England/CAMC-A578EA/2020\|EPI_ISL_597412\|20 | ···················· | ···················· | ···················· | ···················· | ···················· | ···················· |
| N | 4877 | hCoV-19/England/QEUH-A5DDB8/2020\|EPI_ISL_597413\|20 | ···················· | ···················· | ···················· | ···················· | ···················· | ···················· |
| N | 4878 | hCoV-19/England/CAMC-A57C1B/2020\|EPI_ISL_597414\|20 | ···················· | ···················· | ···················· | ···················· | ···················· | ···················· |
| N | 4879 | hCoV-19/England/QEUH-A5E5A0/2020\|EPI_ISL_597415\|20 | ···················· | ···················· | ···················· | ···················· | ···················· | ···················· |
| N | 4880 | hCoV-19/England/QEUH-A5E713/2020\|EPI_ISL_597416\|20 | ···················· | ···················· | ···················· | ···················· | ···················· | ···················· |
| N | 4881 | hCoV-19/England/QEUH-A5E1AC/2020\|EPI_ISL_597417\|20 | ···················· | ···················· | ···················· | ···················· | ···················· | ···················· |
| N | 4882 | hCoV-19/England/CAMC-A57C2A/2020\|EPI_ISL_597418\|20 | ···················· | ···················· | ···················· | ···················· | ···················· | ···················· |
| N | 4883 | hCoV-19/England/QEUH-A5DF58/2020\|EPI_ISL_597419\|20 | ···················· | ···················· | ···················· | ···················· | ···················· | ···················· |
| N | 4884 | hCoV-19/England/QEUH-A5E75F/2020\|EPI_ISL_597422\|20 | ···················· | ···················· | ···················· | ···················· | ···················· | ···················· |
| N | 4885 | hCoV-19/Scotland/QEUH-A5E4B2/2020\|EPI_ISL_597423\|2 | ···················· | ···················· | ···················· | ···················· | ···················· | ···················· |
| N | 4886 | hCoV-19/England/CAMC-A57A7B/2020\|EPI_ISL_597425\|20 | ···················· | ···················· | ···················· | ···················· | ···················· | ···················· |
| N | 4887 | hCoV-19/England/CAMC-A5761D/2020\|EPI_ISL_597426\|20 | ···················· | ···················· | ···················· | ···················· | ···················· | ···················· |
| N | 4888 | hCoV-19/England/CAMC-A57756/2020\|EPI_ISL_597427\|20 | ···················· | ···················· | ···················· | ···················· | ···················· | ···················· |
| N | 4889 | hCoV-19/England/QEUH-A5DEC4/2020\|EPI_ISL_597430\|20 | ···················· | ···················· | ···················· | ···················· | ···················· | ···················· |
| N | 4890 | hCoV-19/England/QEUH-A5E063/2020\|EPI_ISL_597431\|20 | ···················· | ···················· | ···················· | ···················· | ···················· | ···················· |
| N | 4891 | hCoV-19/England/CAMC-A5770B/2020\|EPI_ISL_597432\|20 | ···················· | ···················· | ···················· | ···················· | ···················· | ···················· |
| N | 4892 | hCoV-19/England/MILK-A450AB/2020\|EPI_ISL_598721\|20 | ···················· | ···················· | ···················· | ···················· | ······T············· | ···················· |
| N | 4893 | hCoV-19/England/MILK-A44E48/2020\|EPI_ISL_598722\|20 | ···················· | ···················· | ···················· | ···················· | ···················· | ···················· |
| N | 4894 | hCoV-19/England/MILK-A4668A/2020\|EPI_ISL_598723\|20 | ···················· | ···················· | ···················· | ···················· | ···················· | ···················· |
| N | 4895 | hCoV-19/England/MILK-A45F35/2020\|EPI_ISL_598724\|20 | ···················· | ···················· | ···················· | ···················· | ···················· | ···················· |
| N | 4896 | hCoV-19/England/MILK-A4568B/2020\|EPI_ISL_598725\|20 | ···················· | ···················· | ···················· | ···················· | ···················· | ···················· |
| N | 4897 | hCoV-19/England/MILK-A44EFD/2020\|EPI_ISL_598726\|20 | ···················· | ···················· | ···················· | ···················· | ······T············· | ···················· |
| N | 4898 | hCoV-19/England/MILK-A4576A/2020\|EPI_ISL_598727\|20 | ···················· | ···················· | ···················· | ···················· | ···················· | ···················· |
| N | 4899 | hCoV-19/England/MILK-A44F63/2020\|EPI_ISL_598728\|20 | ···················· | ···················· | ···················· | ···················· | ···················· | ···················· |
| N | 4900 | hCoV-19/England/MILK-A45F26/2020\|EPI_ISL_598729\|20 | ···················· | ···················· | ···················· | ···················· | ···················· | ···················· |
| N | 4901 | hCoV-19/England/MILK-A461F2/2020\|EPI_ISL_598730\|20 | ···················· | ···················· | ···················· | ···················· | ···················· | ···················· |
| N | 4902 | hCoV-19/England/MILK-A45EA1/2020\|EPI_ISL_598731\|20 | ···················· | ···················· | ···················· | ···················· | ···················· | ···················· |
| N | 4903 | hCoV-19/England/MILK-A456D6/2020\|EPI_ISL_598733\|20 | ···················· | ···················· | ···················· | ···················· | ···················· | ···················· |
| N | 4904 | hCoV-19/England/MILK-A44D87/2020\|EPI_ISL_598734\|20 | ···················· | ···················· | ···················· | ···················· | ···················· | ···················· |
| N | 4905 | hCoV-19/England/MILK-A44F72/2020\|EPI_ISL_598735\|20 | ···················· | ···················· | ···················· | ···················· | ···················· | ···················· |

| | | | | | | | | |
|---|---|---|---|---|---|---|---|---|
| N | 4906 | hCoV-19/England/MILK-A4672D/2020\|EPI_ISL_598736\|20 | ···················· | ·A·················· | ···················· | ···················· | ···················· | ···A················ |
| N | 4907 | hCoV-19/England/MILK-A44B23/2020\|EPI_ISL_598737\|20 | ···················· | ···················· | ···················· | ···················· | ···················· | ···················· |
| N | 4908 | hCoV-19/England/MILK-A462D1/2020\|EPI_ISL_598738\|20 | ···················· | ···················· | ···················· | ···················· | ···················· | ···················· |
| N | 4909 | hCoV-19/England/MILK-A458EF/2020\|EPI_ISL_598739\|20 | ···················· | ·A·················· | ···················· | ···················· | ···················· | ···A················ |
| N | 4910 | hCoV-19/England/MILK-A45D1D/2020\|EPI_ISL_598740\|20 | ···················· | ···················· | ···················· | ···················· | ···················· | ···················· |
| N | 4911 | hCoV-19/England/MILK-A44F18/2020\|EPI_ISL_598741\|20 | ···················· | ···················· | ···················· | ···················· | ······T············· | ···················· |
| N | 4912 | hCoV-19/England/MILK-A4615C/2020\|EPI_ISL_598742\|20 | ···················· | ···················· | ···················· | ···················· | ···················· | ···················· |
| N | 4913 | hCoV-19/England/MILK-A46110/2020\|EPI_ISL_598743\|20 | ···················· | ···················· | ···················· | ···················· | ···················· | ···················· |
| N | 4914 | hCoV-19/England/MILK-A44FCD/2020\|EPI_ISL_598747\|20 | ···················· | ···················· | ···················· | ···················· | ···················· | ···················· |
| N | 4915 | hCoV-19/England/MILK-A456B8/2020\|EPI_ISL_598748\|20 | ···················· | ···················· | ···················· | ···················· | ···················· | ···················· |
| N | 4916 | hCoV-19/England/MILK-A44E66/2020\|EPI_ISL_598749\|20 | ···················· | ···················· | ···················· | ···················· | ···················· | ···················· |
| N | 4917 | hCoV-19/England/MILK-A45FF9/2020\|EPI_ISL_598753\|20 | ···················· | ···················· | ···················· | ···················· | ······T············· | ···················· |
| N | 4918 | hCoV-19/England/MILK-A46453/2020\|EPI_ISL_598754\|20 | ···················· | ···················· | ···················· | ···················· | ······T············· | ···················· |
| N | 4919 | hCoV-19/England/MILK-A44ACC/2020\|EPI_ISL_598755\|20 | ···················· | ···················· | ···················· | ···················· | ···················· | ···················· |
| N | 4920 | hCoV-19/England/MILK-A45CF2/2020\|EPI_ISL_598756\|20 | ···················· | ·A·················· | ···················· | ···················· | ···················· | ···A················ |
| N | 4921 | hCoV-19/England/MILK-A4614D/2020\|EPI_ISL_598757\|20 | ···················· | ···················· | ···················· | ···················· | ···················· | ···················· |
| N | 4922 | hCoV-19/England/MILK-A44DD2/2020\|EPI_ISL_598758\|20 | ···················· | ···················· | ···················· | ···················· | ···················· | ···················· |
| N | 4923 | hCoV-19/England/MILK-A45DA4/2020\|EPI_ISL_598760\|20 | ···················· | ···················· | ···················· | ···················· | ······T············· | ···················· |
| N | 4924 | hCoV-19/England/MILK-A45E56/2020\|EPI_ISL_598761\|20 | ···················· | ···················· | ···················· | ···················· | ···················· | ···················· |
| N | 4925 | hCoV-19/England/MILK-A45867/2020\|EPI_ISL_598763\|20 | ···················· | ···················· | ···················· | ···················· | ···················· | ···················· |
| N | 4926 | hCoV-19/England/MILK-A44C3F/2020\|EPI_ISL_598764\|20 | ···················· | ···················· | ···················· | ···················· | ······T············· | ···················· |
| N | 4927 | hCoV-19/Scotland/QEUH-9E4731/2020\|EPI_ISL_601009\|2 | ···················· | ···················· | ···················· | ···················· | ···················· | ···················· |
| N | 4928 | hCoV-19/Scotland/QEUH-9E3EA6/2020\|EPI_ISL_601010\|2 | ···················· | ···················· | ···················· | ···················· | ···················· | ···················· |
| N | 4929 | hCoV-19/Scotland/QEUH-9E4EB4/2020\|EPI_ISL_601011\|2 | ···················· | ·A·················· | ···················· | ···················· | ···················· | ···················· |
| N | 4930 | hCoV-19/Scotland/QEUH-9E54FD/2020\|EPI_ISL_601012\|2 | ···················· | ···················· | ···················· | ···················· | ···················· | ···················· |
| N | 4931 | hCoV-19/Scotland/QEUH-9E4A65/2020\|EPI_ISL_601013\|2 | ···················· | ···················· | ···················· | ···················· | ···················· | ···················· |
| N | 4932 | hCoV-19/Scotland/QEUH-9E41CA/2020\|EPI_ISL_601014\|2 | ···················· | ···················· | ···················· | ···················· | ···················· | ···················· |
| N | 4933 | hCoV-19/Scotland/QEUH-9E5202/2020\|EPI_ISL_601015\|2 | ···················· | ···················· | ···················· | ···················· | ···················· | ···················· |
| N | 4934 | hCoV-19/Scotland/QEUH-9E5299/2020\|EPI_ISL_601016\|2 | ···················· | ······C············· | ···················· | ···················· | ···················· | ···················· |
| N | 4935 | hCoV-19/Scotland/QEUH-9E55EB/2020\|EPI_ISL_601017\|2 | ···················· | ···················· | ···················· | ···················· | ···················· | ···················· |
| N | 4936 | hCoV-19/Scotland/QEUH-9E5590/2020\|EPI_ISL_601018\|2 | ···················· | ···················· | ···················· | ···················· | ···················· | ···················· |
| N | 4937 | hCoV-19/Scotland/QEUH-9E4D20/2020\|EPI_ISL_601019\|2 | ···················· | ···················· | ···················· | ···················· | ···················· | ···················· |
| N | 4938 | hCoV-19/Scotland/QEUH-9E54C0/2020\|EPI_ISL_601020\|2 | ···················· | ···················· | ···················· | ···················· | ···················· | ···················· |
| N | 4939 | hCoV-19/Scotland/QEUH-9E524E/2020\|EPI_ISL_601021\|2 | ···················· | ···················· | ···················· | ···················· | ···················· | ···················· |
| N | 4940 | hCoV-19/Scotland/QEUH-9E42E5/2020\|EPI_ISL_601024\|2 | ···················· | ···················· | ···················· | ···················· | ···················· | ···················· |
| N | 4941 | hCoV-19/England/QEUH-9E53B4/2020\|EPI_ISL_601025\|20 | ···················· | ···················· | ···················· | ···················· | ····T··············· | ···················· |
| N | 4942 | hCoV-19/Scotland/QEUH-9E4090/2020\|EPI_ISL_601026\|2 | ···················· | ···················· | ···················· | ···················· | ···················· | ···················· |
| N | 4943 | hCoV-19/Scotland/QEUH-9E5123/2020\|EPI_ISL_601027\|2 | ···················· | ···················· | ···················· | ···················· | ···················· | ···················· |
| N | 4944 | hCoV-19/Scotland/QEUH-9E51D8/2020\|EPI_ISL_601029\|2 | ···················· | ···················· | ···················· | ···················· | ···················· | ···················· |
| N | 4945 | hCoV-19/Scotland/QEUH-9E5606/2020\|EPI_ISL_601030\|2 | ···················· | ···················· | ···················· | ···················· | ···················· | ···················· |
| N | 4946 | hCoV-19/Scotland/QEUH-9E4564/2020\|EPI_ISL_601031\|2 | ···················· | ···················· | ···················· | ···················· | ···················· | ···················· |
| N | 4947 | hCoV-19/Scotland/QEUH-9E4704/2020\|EPI_ISL_601032\|2 | ···················· | ···················· | ···················· | ···················· | ···················· | ···················· |
| N | 4948 | hCoV-19/Scotland/QEUH-9E542A/2020\|EPI_ISL_601033\|2 | ···················· | ···················· | ···················· | ···················· | ···················· | ···················· |
| N | 4949 | hCoV-19/Scotland/QEUH-9E469E/2020\|EPI_ISL_601034\|2 | ···················· | ···················· | ···················· | ···················· | ···················· | ···················· |
| N | 4950 | hCoV-19/Scotland/QEUH-9E5396/2020\|EPI_ISL_601035\|2 | ···················· | ···················· | ···················· | ···················· | ···················· | ···················· |
| N | 4951 | hCoV-19/Scotland/QEUH-9E42D6/2020\|EPI_ISL_601036\|2 | ···················· | ···················· | ···················· | ···················· | ···················· | ···················· |
| N | 4952 | hCoV-19/Scotland/QEUH-9E43D3/2020\|EPI_ISL_601038\|2 | ···················· | ···················· | ···················· | ···················· | ···················· | ···················· |
| N | 4953 | hCoV-19/Scotland/QEUH-9E4582/2020\|EPI_ISL_601039\|2 | ···················· | ···················· | ···················· | ···················· | ···················· | ···················· |
| N | 4954 | hCoV-19/Scotland/QEUH-9E517E/2020\|EPI_ISL_601040\|2 | ···················· | ···················· | ···················· | ···················· | ···················· | ···················· |
| N | 4955 | hCoV-19/Scotland/QEUH-9E4643/2020\|EPI_ISL_601041\|2 | ···················· | ···················· | ···················· | ···················· | ···················· | ···················· |
| N | 4956 | hCoV-19/Scotland/QEUH-9E435B/2020\|EPI_ISL_601042\|2 | ···················· | ···················· | ···················· | ···················· | ···················· | ···················· |
| N | 4957 | hCoV-19/Scotland/QEUH-9E4D4E/2020\|EPI_ISL_601043\|2 | ···················· | ···················· | ···················· | ···················· | ···················· | ···················· |
| N | 4958 | hCoV-19/Scotland/QEUH-9E46F8/2020\|EPI_ISL_601044\|2 | ···················· | ···················· | ···················· | ···················· | ···················· | ···················· |
| N | 4959 | hCoV-19/Scotland/QEUH-9E482F/2020\|EPI_ISL_601045\|2 | ···················· | ···················· | ···················· | ···················· | ···················· | ···················· |
| N | 4960 | hCoV-19/Scotland/QEUH-9E3F1C/2020\|EPI_ISL_601046\|2 | ···················· | ···················· | ···················· | ···················· | ···················· | ···················· |
| N | 4961 | hCoV-19/Scotland/QEUH-9E4A56/2020\|EPI_ISL_601048\|2 | ···················· | ···················· | ···················· | ···················· | ···················· | ···················· |
| N | 4962 | hCoV-19/Scotland/QEUH-9E44B2/2020\|EPI_ISL_601049\|2 | ···················· | ···················· | ···················· | ···················· | ···················· | ···················· |
| N | 4963 | hCoV-19/Germany/NW-UBI-19/2020\|EPI_ISL_602300\|2020 | ···················· | ···················· | ···················· | ···················· | ···················· | ···················· |
| N | 4964 | hCoV-19/Germany/NW-UBI-20/2020\|EPI_ISL_602301\|2020 | ···················· | ···················· | ···················· | ···················· | ···················· | ···················· |
| N | 4965 | hCoV-19/Germany/NW-UBI-21/2020\|EPI_ISL_602302\|2020 | ···················· | ···················· | ···················· | ···················· | ···················· | ···················· |
| N | 4966 | hCoV-19/Germany/NW-UBI-22/2020\|EPI_ISL_602303\|2020 | ···················· | ···················· | ···················· | ···················· | ···················· | ···················· |
| N | 4967 | hCoV-19/Italy/CAM-IZSM-45946/2020\|EPI_ISL_602304\|2 | ···················· | ···················· | ···················· | ···················· | ···················· | ···················· |
| N | 4968 | hCoV-19/USA/FL-BM3Z/2020\|EPI_ISL_602306\|2020-08-31 | ···················· | ···················· | ···················· | ···················· | ···················· | ···················· |
| N | 4969 | hCoV-19/USA/FL-B2MD/2020\|EPI_ISL_602307\|2020-08-03 | ···················· | ···················· | ···················· | ···················· | ···················· | ···················· |
| N | 4970 | hCoV-19/USA/FL-B2KZ/2020\|EPI_ISL_602308\|2020-08-03 | ···················· | ···················· | ···················· | ···················· | ···················· | ···················· |
| N | 4971 | hCoV-19/USA/FL-BA0B/2020\|EPI_ISL_602309\|2020-08-14 | ···················· | ···················· | ···················· | ···················· | ···················· | ···················· |
| N | 4972 | hCoV-19/USA/FL-B25T/2020\|EPI_ISL_602310\|2020-08-02 | ···················· | ···················· | ···················· | ···················· | ···················· | ···················· |
| N | 4973 | hCoV-19/USA/FL-BBNN/2020\|EPI_ISL_602311\|2020-08-18 | ···················· | ···················· | ···················· | ···················· | ···················· | ···················· |
| N | 4974 | hCoV-19/USA/FL-B2TM/2020\|EPI_ISL_602312\|2020-08-02 | ···················· | ·············A······ | ···················· | ···················· | ···················· | ···················· |
| N | 4975 | hCoV-19/USA/FL-BK0D/2020\|EPI_ISL_602313\|2020-08-28 | ···················· | ·A·················· | ···················· | ···················· | ···················· | ···················· |
| N | 4976 | hCoV-19/USA/FL-B8G5/2020\|EPI_ISL_602314\|2020-08-12 | ···················· | ···················· | ···················· | ···················· | ···················· | ···················· |
| N | 4977 | hCoV-19/USA/FL-B5GJ/2020\|EPI_ISL_602315\|2020-08-05 | ···················· | ···················· | ···················· | ···················· | ···················· | ···················· |
| N | 4978 | hCoV-19/USA/FL-B25U/2020\|EPI_ISL_602316\|2020-08-02 | ···················· | ·············A······ | ···················· | ···················· | ···················· | ···················· |
| N | 4979 | hCoV-19/USA/FL-B2MG/2020\|EPI_ISL_602317\|2020-08-03 | ···················· | ···················· | ···················· | ···················· | ···················· | ···················· |
| N | 4980 | hCoV-19/USA/FL-AZT7/2020\|EPI_ISL_602318\|2020-07-29 | ···················· | ···················· | ···················· | ···················· | ···················· | ···················· |
| N | 4981 | hCoV-19/USA/FL-B5TK/2020\|EPI_ISL_602319\|2020-08-09 | ···················· | ···················· | ···················· | ···················· | ···················· | ···················· |
| N | 4982 | hCoV-19/USA/FL-B4R9/2020\|EPI_ISL_602322\|2020-08-05 | ···················· | ···················· | ···················· | ···················· | ···················· | ···················· |
| N | 4983 | hCoV-19/USA/FL-B801/2020\|EPI_ISL_602323\|2020-08-12 | ···················· | ···················· | ···················· | ···················· | ···················· | ···················· |
| N | 4984 | hCoV-19/USA/PR-B2D2/2020\|EPI_ISL_602325\|2020-08-03 | ···················· | ···················· | ···················· | ···················· | ···················· | ···················· |
| N | 4985 | hCoV-19/USA/PR-B2D3/2020\|EPI_ISL_602326\|2020-08-02 | ···················· | ···················· | ···················· | ···················· | ···················· | ···················· |

| | | | | | | | | |
|---|---|---|---|---|---|---|---|---|
| N | 4986 hCoV-19/USA/PR-B2F0/2020\|EPI_ISL_602327\|2020-08-03 | ···················· | ···················· | ···················· | ···················· | ···················· | ···················· | |
| N | 4987 hCoV-19/USA/PR-B2F8/2020\|EPI_ISL_602328\|2020-08-01 | ···················· | ···················· | ···················· | ···················· | ···················· | ···················· | |
| N | 4988 hCoV-19/Russia/SPE-RII-MH1598S/2020\|EPI_ISL_602340 | ···················· | ···················· | ···················· | ···················· | ···················· | ···················· | |
| N | 4989 hCoV-19/Russia/SPE-RII-MH1624S/2020\|EPI_ISL_602341 | ···················· | ···················· | ···················· | ···················· | ······T············· | ···················· | |
| N | 4990 hCoV-19/Russia/SVE-RII-MH1691S/2020\|EPI_ISL_602342 | ···················· | ···················· | ···················· | ···················· | ···················· | ···················· | |
| N | 4991 hCoV-19/Russia/SVE-RII-MH1697S/2020\|EPI_ISL_602343 | ···················· | ···················· | ···················· | ···················· | ···················· | ···················· | |
| N | 4992 hCoV-19/Russia/MUR-RII-MH1700S/2020\|EPI_ISL_602344 | ···················· | ·A·················· | ···················· | ···················· | ···················· | ···················· | |
| N | 4993 hCoV-19/Russia/MUR-RII-MH1701S/2020\|EPI_ISL_602345 | ···················· | ·A·················· | ···················· | ···················· | ···················· | ···················· | |
| N | 4994 hCoV-19/Russia/NGR-RII-MH1702S/2020\|EPI_ISL_602346 | ···················· | ···················· | ···················· | ···················· | ···················· | ···················· | |
| N | 4995 hCoV-19/Russia/NGR-RII-MH1705S/2020\|EPI_ISL_602347 | ···················· | ···················· | ···················· | ···················· | ···················· | ···················· | |
| N | 4996 hCoV-19/Russia/SVE-RII-MH1711S/2020\|EPI_ISL_602348 | ···················· | ···················· | ···················· | ···················· | ···················· | ···················· | |
| N | 4997 hCoV-19/Russia/SVE-RII-MH1712S/2020\|EPI_ISL_602349 | ···················· | ···················· | ···················· | ···················· | ···················· | ···················· | |
| N | 4998 hCoV-19/Russia/SVE-RII-MH1713S/2020\|EPI_ISL_602350 | ···················· | ···················· | ···················· | ···················· | ···················· | ···················· | |
| N | 4999 hCoV-19/Russia/SVE-RII-MH1714S/2020\|EPI_ISL_602351 | ···················· | ···················· | ···················· | ···················· | ···················· | ···················· | |
| N | 5000 hCoV-19/Russia/SVE-RII-MH1715S/2020\|EPI_ISL_602352 | ···················· | ···················· | ···················· | ···················· | ···················· | ···················· | |
| N | 5001 hCoV-19/Russia/SVE-RII-MH1716S/2020\|EPI_ISL_602353 | ···················· | ···················· | ···················· | ···················· | ···················· | ···················· | |
| N | 5002 hCoV-19/Russia/SVE-RII-MH1717S/2020\|EPI_ISL_602354 | ···················· | ···················· | ···················· | ···················· | ···················· | ···················· | |
| N | 5003 hCoV-19/Russia/SVE-RII-MH1720S/2020\|EPI_ISL_602355 | ···················· | ···················· | ···················· | ···················· | ···················· | ···················· | |
| N | 5004 hCoV-19/Russia/SVE-RII-MH1722S/2020\|EPI_ISL_602356 | ···················· | ···················· | ···················· | ···················· | ···················· | ···················· | |
| N | 5005 hCoV-19/Russia/SVE-RII-MH1723S/2020\|EPI_ISL_602357 | ···················· | ···················· | ···················· | ···················· | ···················· | ···················· | |
| N | 5006 hCoV-19/Russia/SVE-RII-MH1724S/2020\|EPI_ISL_602358 | ···················· | ···················· | ···················· | ···················· | ···················· | ···················· | |
| N | 5007 hCoV-19/Russia/SVE-RII-MH1725S/2020\|EPI_ISL_602359 | ···················· | ···················· | ···················· | ···················· | ···················· | ···················· | |
| N | 5008 hCoV-19/Russia/SVE-RII-MH1727S/2020\|EPI_ISL_602360 | ···················· | ···················· | ···················· | ···················· | ···················· | ···················· | |
| N | 5009 hCoV-19/Russia/SVE-RII-MH1733S/2020\|EPI_ISL_602361 | ···················· | ···················· | ···················· | ···················· | ···················· | ···················· | |
| N | 5010 hCoV-19/Russia/SVE-RII-MH1758S/2020\|EPI_ISL_602362 | ···················· | ···················· | ···················· | ···················· | ···················· | ···················· | |
| N | 5011 hCoV-19/Russia/SVE-RII-MH1910S/2020\|EPI_ISL_602363 | ···················· | ···················· | ···················· | ···················· | ···················· | ···················· | |
| N | 5012 hCoV-19/Russia/KHM-RII-MH1914S/2020\|EPI_ISL_602364 | ···················· | ···················· | ···················· | ···················· | ···················· | ···················· | |
| N | 5013 hCoV-19/Russia/PER-RII-MH1915S/2020\|EPI_ISL_602365 | ···················· | ···················· | ···················· | ···················· | ···················· | ···················· | |
| N | 5014 hCoV-19/Russia/SVE-RII-MH1918S/2020\|EPI_ISL_602366 | ···················· | ···················· | ···················· | ···················· | ·················C·· | ···················· | 1 |
| N | 5015 hCoV-19/Russia/SVE-RII-MH1924S/2020\|EPI_ISL_602367 | ···················· | ···················· | ···················· | ···················· | ···················· | ···················· | |
| N | 5016 hCoV-19/Russia/SVE-RII-MH1927S/2020\|EPI_ISL_602368 | ···················· | ···················· | ···················· | ···················· | ···················· | ···················· | |
| N | 5017 hCoV-19/Russia/SVE-RII-MH1928S/2020\|EPI_ISL_602369 | ···················· | ···················· | ···················· | ···················· | ···················· | ···················· | |
| N | 5018 hCoV-19/Russia/SVE-RII-MH1930S/2020\|EPI_ISL_602370 | ···················· | ···················· | ···················· | ···················· | ···················· | ···················· | |
| N | 5019 hCoV-19/Russia/SVE-RII-MH1931S/2020\|EPI_ISL_602371 | ···················· | ···················· | ···················· | ···················· | ···················· | ···················· | |
| N | 5020 hCoV-19/Russia/SVE-RII-MH1932S/2020\|EPI_ISL_602372 | ···················· | ···················· | ···················· | ···················· | ···················· | ···················· | |
| N | 5021 hCoV-19/Russia/SVE-RII-MH1936S/2020\|EPI_ISL_602373 | ···················· | ···················· | ···················· | ···················· | ·········A·········· | ···················· | |
| N | 5022 hCoV-19/Russia/SVE-RII-MH1941S/2020\|EPI_ISL_602374 | ···················· | ···················· | ···················· | ···················· | ···················· | ···················· | |
| N | 5023 hCoV-19/Russia/PER-RII-MH1950S/2020\|EPI_ISL_602375 | ···················· | ···················· | ···················· | ···················· | ···················· | ···················· | |
| N | 5024 hCoV-19/Russia/PER-RII-MH1951S/2020\|EPI_ISL_602376 | ···················· | ···················· | ···················· | ···················· | ···················· | ···················· | |
| N | 5025 hCoV-19/Russia/PSK-RII-MH2017S/2020\|EPI_ISL_602377 | ···················· | ···················· | ···················· | ···················· | ···················· | ···················· | |
| N | 5026 hCoV-19/Russia/PSK-RII-MH2018S/2020\|EPI_ISL_602378 | ···················· | ···················· | ···················· | ···················· | ···················· | ···················· | |
| N | 5027 hCoV-19/Russia/NGR-RII-MH2019S/2020\|EPI_ISL_602379 | ···················· | ···················· | ···················· | ···················· | ···················· | ···················· | |
| N | 5028 hCoV-19/Switzerland/VD-ETHZ-321384/2020\|EPI_ISL_60 | ···················· | ···················· | ···················· | ···················· | ···················· | ···················· | |
| N | 5029 hCoV-19/Switzerland/UR-ETHZ-321385/2020\|EPI_ISL_60 | ···················· | ···················· | ···················· | ···················· | ···················· | ···················· | |
| N | 5030 hCoV-19/Switzerland/VD-ETHZ-321387/2020\|EPI_ISL_60 | ···················· | ···················· | ···················· | ···················· | ···················· | ···················· | |
| N | 5031 hCoV-19/Switzerland/VD-ETHZ-321388/2020\|EPI_ISL_60 | ···················· | ···················· | ···················· | ···················· | ···················· | ···················· | |
| N | 5032 hCoV-19/Switzerland/VS-ETHZ-321390/2020\|EPI_ISL_60 | ···················· | ···················· | ···················· | ···················· | ···················· | ···················· | |
| N | 5033 hCoV-19/Switzerland/VD-ETHZ-321391/2020\|EPI_ISL_60 | ···················· | ···················· | ···················· | ···················· | ···················· | ···················· | |
| N | 5034 hCoV-19/Switzerland/VD-ETHZ-321392/2020\|EPI_ISL_60 | ···················· | ···················· | ···················· | ···················· | ···················· | ···················· | |
| N | 5035 hCoV-19/Switzerland/VD-ETHZ-321393/2020\|EPI_ISL_60 | ···················· | ···················· | ···················· | ···················· | ···················· | ···················· | |
| N | 5036 hCoV-19/Switzerland/VD-ETHZ-321394/2020\|EPI_ISL_60 | ···················· | ···················· | ···················· | ···················· | ···················· | ···················· | |
| N | 5037 hCoV-19/Switzerland/TI-ETHZ-321395/2020\|EPI_ISL_60 | ···················· | ···················· | ···················· | ···················· | ···················· | ···················· | |
| N | 5038 hCoV-19/Switzerland/BE-ETHZ-321396/2020\|EPI_ISL_60 | ···················· | ···················· | ···················· | ···················· | ······K············· | ···················· | |
| N | 5039 hCoV-19/Switzerland/BE-ETHZ-321397/2020\|EPI_ISL_60 | ···················· | ···················· | ···················· | ···················· | ···················· | ···················· | |
| N | 5040 hCoV-19/Switzerland/VD-ETHZ-321398/2020\|EPI_ISL_60 | ···················· | ···················· | ···················· | ···················· | ···················· | ···················· | |
| N | 5041 hCoV-19/Switzerland/VD-ETHZ-321399/2020\|EPI_ISL_60 | ···················· | ···················· | ···················· | ···················· | ···················· | ···················· | |
| N | 5042 hCoV-19/Switzerland/JU-ETHZ-321400/2020\|EPI_ISL_60 | ···················· | ···················· | ···················· | ···················· | ···················· | ···················· | |
| N | 5043 hCoV-19/Switzerland/VS-ETHZ-321401/2020\|EPI_ISL_60 | ···················· | ···················· | ···················· | ···················· | ···················· | ···················· | |
| N | 5044 hCoV-19/Switzerland/VS-ETHZ-321402/2020\|EPI_ISL_60 | ···················· | ···················· | ···················· | ···················· | ···················· | ···················· | |
| N | 5045 hCoV-19/Switzerland/TG-ETHZ-321403/2020\|EPI_ISL_60 | ···················· | ···················· | ···················· | ···················· | ···················· | ···················· | |
| N | 5046 hCoV-19/Switzerland/VS-ETHZ-321404/2020\|EPI_ISL_60 | ···················· | ···················· | ···················· | ···················· | ···················· | ···················· | |
| N | 5047 hCoV-19/Switzerland/VS-ETHZ-321405/2020\|EPI_ISL_60 | ···················· | ···················· | ···················· | ···················· | ···················· | ···················· | |
| N | 5048 hCoV-19/Switzerland/BE-ETHZ-321406/2020\|EPI_ISL_60 | ···················· | ···················· | ···················· | ···················· | ···················· | ···················· | |
| N | 5049 hCoV-19/Switzerland/BE-ETHZ-321407/2020\|EPI_ISL_60 | ···················· | ···················· | ···················· | ···················· | ···················· | ···················· | |
| N | 5050 hCoV-19/Switzerland/VD-ETHZ-321408/2020\|EPI_ISL_60 | ···················· | ···················· | ···················· | ···················· | ···················· | ···················· | |
| N | 5051 hCoV-19/Switzerland/BL-ETHZ-321412/2020\|EPI_ISL_60 | ···················· | ···················· | ···················· | ···················· | ···················· | ···················· | |
| N | 5052 hCoV-19/Switzerland/BE-ETHZ-321413/2020\|EPI_ISL_60 | ···················· | ···················· | ···················· | ···················· | ···················· | ···················· | |
| N | 5053 hCoV-19/Switzerland/BL-ETHZ-321414/2020\|EPI_ISL_60 | ···················· | ···················· | ···················· | ···················· | ···················· | ···················· | |
| N | 5054 hCoV-19/Switzerland/BL-ETHZ-321416/2020\|EPI_ISL_60 | ···················· | ···················· | ···················· | ···················· | ···················· | ···················· | |
| N | 5055 hCoV-19/Switzerland/VS-ETHZ-321417/2020\|EPI_ISL_60 | ···················· | ···················· | ···················· | ···················· | ···················· | ···················· | |
| N | 5056 hCoV-19/Switzerland/BL-ETHZ-321418/2020\|EPI_ISL_60 | ···················· | ···················· | ···················· | ···················· | ······K············· | ···················· | |
| N | 5057 hCoV-19/Switzerland/ZH-ETHZ-321420/2020\|EPI_ISL_60 | ···················· | ···················· | ···················· | ···················· | ······K············· | ···················· | |
| N | 5058 hCoV-19/Switzerland/VD-ETHZ-321421/2020\|EPI_ISL_60 | ···················· | ···················· | ···················· | ···················· | ···················· | ···················· | |
| N | 5059 hCoV-19/Switzerland/VD-ETHZ-321447/2020\|EPI_ISL_60 | ···················· | ···················· | ···················· | ···················· | ···················· | ···················· | |
| N | 5060 hCoV-19/Switzerland/TI-ETHZ-321449/2020\|EPI_ISL_60 | ···················· | ···················· | ···················· | ···················· | ···················· | ···················· | |
| N | 5061 hCoV-19/Switzerland/UR-ETHZ-321450/2020\|EPI_ISL_60 | ···················· | ···················· | ···················· | ···················· | ···················· | ···················· | |
| N | 5062 hCoV-19/Switzerland/UR-ETHZ-321458/2020\|EPI_ISL_60 | ···················· | ···················· | ···················· | ···················· | ···················· | ···················· | |
| N | 5063 hCoV-19/Switzerland/UR-ETHZ-321459/2020\|EPI_ISL_60 | ···················· | ···················· | ···················· | ···················· | ···················· | ···················· | |
| N | 5064 hCoV-19/Switzerland/VS-ETHZ-321460/2020\|EPI_ISL_60 | ···················· | ···················· | ···················· | ···················· | ···················· | ···················· | |
| N | 5065 hCoV-19/Switzerland/BL-ETHZ-321461/2020\|EPI_ISL_60 | ···················· | ···················· | ···················· | ···················· | ······K············· | ···················· | |

| | | | | | | | | |
|---|---|---|---|---|---|---|---|---|
| N | 5066 | hCoV-19/Switzerland/VD-ETHZ-321462/2020\|EPI_ISL_60 | ···················· | ···················· | ···················· | ···················· | ···················· | ···················· |
| N | 5067 | hCoV-19/Switzerland/VD-ETHZ-321463/2020\|EPI_ISL_60 | ···················· | ···················· | ···················· | ···················· | ···················· | ···················· |
| N | 5068 | hCoV-19/Switzerland/GR-ETHZ-321464/2020\|EPI_ISL_60 | ···················· | ···················· | ···················· | ···················· | ···················· | ···················· |
| N | 5069 | hCoV-19/Switzerland/VD-ETHZ-321465/2020\|EPI_ISL_60 | ···················· | ···················· | ···················· | ···················· | ···················· | ···················· |
| N | 5070 | hCoV-19/Switzerland/VD-ETHZ-321467/2020\|EPI_ISL_60 | ···················· | ···················· | ···················· | ···················· | ···················· | ···················· |
| N | 5071 | hCoV-19/Switzerland/BS-ETHZ-321468/2020\|EPI_ISL_60 | ···················· | ·A·················· | ···················· | ···················· | ···················· | ···················· |
| N | 5072 | hCoV-19/Switzerland/VD-ETHZ-321470/2020\|EPI_ISL_60 | ···················· | ···················· | ···················· | ···················· | ···················· | ···················· |
| N | 5073 | hCoV-19/Switzerland/GE-ETHZ-321471/2020\|EPI_ISL_60 | ···················· | ···················· | ···················· | ···················· | ···················· | ···················· |
| N | 5074 | hCoV-19/Switzerland/GE-ETHZ-321472/2020\|EPI_ISL_60 | ···················· | ···················· | ···················· | ···················· | ···················· | ···················· |
| N | 5075 | hCoV-19/Switzerland/GE-ETHZ-321473/2020\|EPI_ISL_60 | ···················· | ···················· | ···················· | ···················· | ···················· | ···················· |
| N | 5076 | hCoV-19/Switzerland/GE-ETHZ-321475/2020\|EPI_ISL_60 | ···················· | ···················· | ···················· | ···················· | ···················· | ···················· |
| N | 5077 | hCoV-19/Switzerland/BE-ETHZ-321476/2020\|EPI_ISL_60 | ···················· | ···················· | ···················· | ···················· | ···················· | ···················· |
| N | 5078 | hCoV-19/Switzerland/VD-ETHZ-321477/2020\|EPI_ISL_60 | ···················· | ···················· | ···················· | ···················· | ···················· | ···················· |
| N | 5079 | hCoV-19/Switzerland/VS-ETHZ-321478/2020\|EPI_ISL_60 | ···················· | ···················· | ···················· | ···················· | ···················· | ···················· |
| N | 5080 | hCoV-19/Switzerland/VS-ETHZ-321479/2020\|EPI_ISL_60 | ···················· | ···················· | ···················· | ···················· | ···················· | ···················· |
| N | 5081 | hCoV-19/Switzerland/GE-ETHZ-321480/2020\|EPI_ISL_60 | ···················· | ···················· | ···················· | ···················· | ···················· | ···················· |
| N | 5082 | hCoV-19/Switzerland/BE-ETHZ-321481/2020\|EPI_ISL_60 | ···················· | ···················· | ···················· | ···················· | ···················· | ···················· |
| N | 5083 | hCoV-19/Switzerland/VS-ETHZ-321482/2020\|EPI_ISL_60 | ···················· | ···················· | ···················· | ···················· | ···················· | ···················· |
| N | 5084 | hCoV-19/Switzerland/VS-ETHZ-321483/2020\|EPI_ISL_60 | ···················· | ···················· | ···················· | ···················· | ···················· | ···················· |
| N | 5085 | hCoV-19/Switzerland/GE-ETHZ-321484/2020\|EPI_ISL_60 | ···················· | ···················· | ···················· | ···················· | ···················· | ···················· |
| N | 5086 | hCoV-19/Switzerland/VD-ETHZ-321485/2020\|EPI_ISL_60 | ···················· | ···················· | ···················· | ···················· | ···················· | ···················· |
| N | 5087 | hCoV-19/Switzerland/VD-ETHZ-321486/2020\|EPI_ISL_60 | ···················· | ···················· | ···················· | ···················· | ···················· | ···················· |
| N | 5088 | hCoV-19/Switzerland/BE-ETHZ-321487/2020\|EPI_ISL_60 | ···················· | ···················· | ···················· | ···················· | ···················· | ···················· |
| N | 5089 | hCoV-19/Switzerland/BE-ETHZ-321488/2020\|EPI_ISL_60 | ···················· | ···················· | ···················· | ···················· | ···················· | ···················· |
| N | 5090 | hCoV-19/Switzerland/BE-ETHZ-321490/2020\|EPI_ISL_60 | ···················· | ···················· | ···················· | ···················· | ···················· | ···················· |
| N | 5091 | hCoV-19/Switzerland/BE-ETHZ-321491/2020\|EPI_ISL_60 | ···················· | ···················· | ···················· | ···················· | ······K············· | ···················· |
| N | 5092 | hCoV-19/USA/NY-QDX-2667/2020\|EPI_ISL_604958\|2020-0 | ···················· | ···················· | ···················· | ···················· | ···················· | ···················· |
| N | 5093 | hCoV-19/USA/OH-QDX-2655/2020\|EPI_ISL_604959\|2020-0 | ···················· | ···················· | ···················· | ···················· | ···················· | ···················· |
| N | 5094 | hCoV-19/USA/PA-QDX-2606/2020\|EPI_ISL_604960\|2020-0 | ···················· | ···················· | ···················· | ···················· | ···················· | ···················· |
| N | 5095 | hCoV-19/USA/UT-QDX-2569/2020\|EPI_ISL_604961\|2020-0 | ···················· | ···················· | ···················· | ···················· | ···················· | ···················· |
| N | 5096 | hCoV-19/USA/UT-QDX-2572/2020\|EPI_ISL_604962\|2020-0 | ···················· | ···················· | ···················· | ···················· | ···················· | ···················· |
| N | 5097 | hCoV-19/USA/UT-QDX-2575/2020\|EPI_ISL_604963\|2020-0 | ···················· | ···················· | ···················· | ···················· | ···················· | ···················· |
| N | 5098 | hCoV-19/USA/UT-QDX-2580/2020\|EPI_ISL_604964\|2020-0 | ···················· | ···················· | ···················· | ···················· | ···················· | ···················· |
| N | 5099 | hCoV-19/USA/UT-QDX-2589/2020\|EPI_ISL_604965\|2020-0 | ···················· | ···················· | ···················· | ···················· | ···················· | ···················· |
| N | 5100 | hCoV-19/USA/UT-QDX-2596/2020\|EPI_ISL_604966\|2020-0 | ···················· | ···················· | ···················· | ···················· | ···················· | ···················· |
| N | 5101 | hCoV-19/USA/UT-QDX-2598/2020\|EPI_ISL_604967\|2020-0 | ···················· | ···················· | ···················· | ···················· | ···················· | ···················· |
| N | 5102 | hCoV-19/USA/UT-QDX-2599/2020\|EPI_ISL_604968\|2020-0 | ···················· | ···················· | ···················· | ···················· | ···················· | ···················· |
| N | 5103 | hCoV-19/USA/UT-QDX-2607/2020\|EPI_ISL_604969\|2020-0 | ···················· | ···················· | ···················· | ···················· | ···················· | ···················· |
| N | 5104 | hCoV-19/USA/UT-QDX-2614/2020\|EPI_ISL_604970\|2020-0 | ···················· | ···················· | ···················· | ···················· | ···················· | ···················· |
| N | 5105 | hCoV-19/USA/UT-QDX-2621/2020\|EPI_ISL_604971\|2020-0 | ···················· | ···················· | ···················· | ···················· | ···················· | ···················· |
| N | 5106 | hCoV-19/USA/UT-QDX-2626/2020\|EPI_ISL_604972\|2020-0 | ···················· | ···················· | ···················· | ···················· | ···················· | ···················· |
| N | 5107 | hCoV-19/USA/UT-QDX-2628/2020\|EPI_ISL_604973\|2020-0 | ···················· | ···················· | ···················· | ···················· | ···················· | ···················· |
| N | 5108 | hCoV-19/USA/UT-QDX-2632/2020\|EPI_ISL_604974\|2020-0 | ···················· | ···················· | ···················· | ···················· | ···················· | ···················· |
| N | 5109 | hCoV-19/USA/UT-QDX-2634/2020\|EPI_ISL_604975\|2020-0 | ···················· | ···················· | ···················· | ···················· | ···················· | ···················· |
| N | 5110 | hCoV-19/USA/UT-QDX-2638/2020\|EPI_ISL_604976\|2020-0 | ···················· | ···················· | ···················· | ···················· | ···················· | ···················· |
| N | 5111 | hCoV-19/USA/UT-QDX-2643/2020\|EPI_ISL_604977\|2020-0 | ···················· | ···················· | ···················· | ···················· | ···················· | ···················· |
| N | 5112 | hCoV-19/USA/UT-QDX-2644/2020\|EPI_ISL_604978\|2020-0 | ···················· | ···················· | ···················· | ···················· | ···················· | ···················· |
| N | 5113 | hCoV-19/USA/UT-QDX-2649/2020\|EPI_ISL_604979\|2020-0 | ···················· | ···················· | ···················· | ···················· | ···················· | ···················· |
| N | 5114 | hCoV-19/USA/UT-QDX-2657/2020\|EPI_ISL_604983\|2020-0 | ···················· | ···················· | ···················· | ···················· | ···················· | ···················· |
| N | 5115 | hCoV-19/USA/UT-QDX-2663/2020\|EPI_ISL_604984\|2020-0 | ···················· | ···················· | ···················· | ···················· | ···················· | ···················· |
| N | 5116 | hCoV-19/USA/UT-QDX-2664/2020\|EPI_ISL_604985\|2020-0 | ···················· | ···················· | ···················· | ···················· | ···················· | ···················· |
| N | 5117 | hCoV-19/USA/UT-QDX-2668/2020\|EPI_ISL_604986\|2020-0 | ···················· | ···················· | ···················· | ···················· | ···················· | ···················· |
| N | 5118 | hCoV-19/USA/UT-QDX-2669/2020\|EPI_ISL_604987\|2020-0 | ···················· | ···················· | ···················· | ···················· | ···················· | ···················· |
| N | 5119 | hCoV-19/USA/UT-QDX-2671/2020\|EPI_ISL_604988\|2020-0 | ···················· | ···················· | ···················· | ···················· | ···················· | ···················· |
| N | 5120 | hCoV-19/USA/UT-QDX-2672/2020\|EPI_ISL_604989\|2020-0 | ···················· | ···················· | ···················· | ···················· | ···················· | ···················· |
| N | 5121 | hCoV-19/England/QEUH-A5E2D6/2020\|EPI_ISL_597280\|20 | ···················· | ···················· | ···················· | ···················· | ···················· | ···················· |
| N | 5122 | hCoV-19/England/QEUH-A5E133/2020\|EPI_ISL_597281\|20 | ···················· | ···················· | ···················· | ···················· | ···················· | ···················· |
| N | 5123 | hCoV-19/England/CAMC-A578AE/2020\|EPI_ISL_597282\|20 | ···················· | ···················· | ···················· | ···················· | ···················· | ···················· |
| N | 5124 | hCoV-19/England/CAMC-A57BF0/2020\|EPI_ISL_597283\|20 | ···················· | ···················· | ···················· | ···················· | ···················· | ···················· |
| N | 5125 | hCoV-19/England/CAMC-A57A4E/2020\|EPI_ISL_597284\|20 | ···················· | ···················· | ···················· | ···················· | ······T············· | ···················· |
| N | 5126 | hCoV-19/England/CAMC-A57CA2/2020\|EPI_ISL_597285\|20 | ···················· | ···················· | ···················· | ···················· | ···················· | ···················· |
| N | 5127 | hCoV-19/England/CAMC-A5762C/2020\|EPI_ISL_597286\|20 | ···················· | ···················· | ···················· | ···················· | ···················· | ···················· |
| N | 5128 | hCoV-19/England/CAMC-A56F7D/2020\|EPI_ISL_597287\|20 | ···················· | ···················· | ···················· | ···················· | ···················· | ···················· |
| N | 5129 | hCoV-19/England/QEUH-A5DE2E/2020\|EPI_ISL_597288\|20 | ···················· | ·A·················· | ···················· | ···················· | ···················· | ··A················· |
| N | 5130 | hCoV-19/England/CAMC-A5702E/2020\|EPI_ISL_597289\|20 | ···················· | ···················· | ···················· | ···················· | ···················· | ···················· |
| N | 5131 | hCoV-19/England/QEUH-A5DE00/2020\|EPI_ISL_597290\|20 | ···················· | ···················· | ···················· | ···················· | ···················· | ···················· |
| N | 5132 | hCoV-19/England/QEUH-A5E1F7/2020\|EPI_ISL_597291\|20 | ···················· | ···················· | ···················· | ···················· | ···················· | ···················· |
| N | 5133 | hCoV-19/England/QEUH-A5E94A/2020\|EPI_ISL_597292\|20 | ···················· | ···················· | ···················· | ···················· | ···················· | ···················· |
| N | 5134 | hCoV-19/England/CAMC-A577B0/2020\|EPI_ISL_597293\|20 | ···················· | ···················· | ···················· | ···················· | ···················· | ···················· |
| N | 5135 | hCoV-19/England/CAMC-A575B6/2020\|EPI_ISL_597295\|20 | ···················· | ···················· | ···················· | ···················· | ···················· | ···················· |
| N | 5136 | hCoV-19/England/QEUH-A5E33D/2020\|EPI_ISL_597296\|20 | ···················· | ···················· | ···················· | ···················· | ···················· | ···················· |
| N | 5137 | hCoV-19/England/CAMC-A57C48/2020\|EPI_ISL_597297\|20 | ···················· | ···················· | ···················· | ···················· | ···················· | ···················· |
| N | 5138 | hCoV-19/England/QEUH-A5E6E9/2020\|EPI_ISL_597298\|20 | ···················· | ···················· | ···················· | ···················· | ···················· | ···················· |
| N | 5139 | hCoV-19/England/QEUH-A5E50A/2020\|EPI_ISL_597299\|20 | ···················· | ···················· | ···················· | ···················· | ···················· | ···················· |
| N | 5140 | hCoV-19/England/QEUH-A5E607/2020\|EPI_ISL_597314\|20 | ···················· | ···················· | ···················· | ···················· | ···················· | ···················· |
| N | 5141 | hCoV-19/England/QEUH-A5E081/2020\|EPI_ISL_597315\|20 | ···················· | ···················· | ···················· | ···················· | ···················· | ···················· |
| N | 5142 | hCoV-19/England/CAMC-A57000/2020\|EPI_ISL_597316\|20 | ···················· | ···················· | ···················· | ···················· | ···················· | ···················· |
| N | 5143 | hCoV-19/England/QEUH-A5E76E/2020\|EPI_ISL_597318\|20 | ···················· | ···················· | ···················· | ···················· | ···················· | ···················· |
| N | 5144 | hCoV-19/England/CAMC-A57370/2020\|EPI_ISL_597319\|20 | ···················· | ···················· | ···················· | ···················· | ······T············· | ···················· |
| N | 5145 | hCoV-19/England/CAMC-A57BE1/2020\|EPI_ISL_597320\|20 | ···················· | ···················· | ···················· | ···················· | ···················· | ···················· |

| | | | | | | | | |
|---|---|---|---|---|---|---|---|---|
| N | 5146 | hCoV-19/England/CAMC-A57C0C/2020\|EPI_ISL_597321\|20 | ···················· | ···················· | ···················· | ···················· | ···················· | ···················· |
| N | 5147 | hCoV-19/England/QEUH-A5E3E2/2020\|EPI_ISL_597322\|20 | ···················· | ···················· | ···················· | ···················· | ···················· | ···················· |
| N | 5148 | hCoV-19/England/CAMC-A573BC/2020\|EPI_ISL_597323\|20 | ···················· | ···················· | ···················· | ···················· | ···················· | ···················· |
| N | 5149 | hCoV-19/England/CAMC-A572A0/2020\|EPI_ISL_597324\|20 | ···················· | ···················· | ···················· | ···················· | ···················· | ···················· |
| N | 5150 | hCoV-19/England/CAMC-A57BA5/2020\|EPI_ISL_597325\|20 | ···················· | ···················· | ···················· | ···················· | ···················· | ···················· |
| N | 5151 | hCoV-19/England/QEUH-A5E7F5/2020\|EPI_ISL_597326\|20 | ···················· | ···················· | ···················· | ···················· | ···················· | ···················· |
| N | 5152 | hCoV-19/England/CAMC-A57422/2020\|EPI_ISL_597327\|20 | ···················· | ···················· | ···················· | ···················· | ···················· | ···················· |
| N | 5153 | hCoV-19/England/QEUH-A5E6DA/2020\|EPI_ISL_597328\|20 | ···················· | ···················· | ···················· | ···················· | ···················· | ···················· |
| N | 5154 | hCoV-19/England/QEUH-A5E68F/2020\|EPI_ISL_597329\|20 | ···················· | ···················· | ···················· | ···················· | ···················· | ···················· |
| N | 5155 | hCoV-19/England/QEUH-A5DEA6/2020\|EPI_ISL_597330\|20 | ···················· | ···················· | ···················· | ···················· | ···················· | ···················· |
| N | 5156 | hCoV-19/England/CAMC-A57361/2020\|EPI_ISL_597331\|20 | ···················· | ···················· | ···················· | ···················· | ···················· | ···················· |
| N | 5157 | hCoV-19/England/QEUH-A5E801/2020\|EPI_ISL_597332\|20 | ···················· | ···················· | ···················· | ···················· | ···················· | ···················· |
| N | 5158 | hCoV-19/England/QEUH-A5DE88/2020\|EPI_ISL_597333\|20 | ···················· | ···················· | ···················· | ···················· | ···················· | ···················· |
| N | 5159 | hCoV-19/England/QEUH-A5E8F2/2020\|EPI_ISL_597334\|20 | ···················· | ···················· | ···················· | ···················· | ···················· | ···················· |
| N | 5160 | hCoV-19/England/CAMC-A56FC8/2020\|EPI_ISL_597335\|20 | ···················· | ···················· | ···················· | ···················· | ···················· | ···················· |
| N | 5161 | hCoV-19/England/QEUH-A5E959/2020\|EPI_ISL_597336\|20 | ···················· | ···················· | ···················· | ···················· | ···················· | ···················· |
| N | 5162 | hCoV-19/England/CAMC-A5752F/2020\|EPI_ISL_597337\|20 | ···················· | ···················· | ···················· | ···················· | ···················· | ···················· |
| N | 5163 | hCoV-19/England/QEUH-A5DD8B/2020\|EPI_ISL_597338\|20 | ···················· | ···················· | ···················· | ···················· | ···················· | ···················· |
| N | 5164 | hCoV-19/England/QEUH-A5E5BF/2020\|EPI_ISL_597339\|20 | ···················· | ···················· | ···················· | ···················· | ···················· | ···················· |
| N | 5165 | hCoV-19/England/CAMC-A57A5D/2020\|EPI_ISL_597340\|20 | ···················· | ···················· | ···················· | ···················· | ···················· | ···················· |
| N | 5166 | hCoV-19/England/CAMC-A577ED/2020\|EPI_ISL_597342\|20 | ···················· | ···················· | ···················· | ···················· | ···················· | ···················· |
| N | 5167 | hCoV-19/England/CAMC-A5738F/2020\|EPI_ISL_597343\|20 | ···················· | ···················· | ···················· | ···················· | ···················· | ···················· |
| N | 5168 | hCoV-19/England/CAMC-A57AD5/2020\|EPI_ISL_597345\|20 | ···················· | ···················· | ···················· | ···················· | ···················· | ···················· |
| N | 5169 | hCoV-19/England/CAMC-A57914/2020\|EPI_ISL_597346\|20 | ···················· | ···················· | ···················· | ···················· | ···················· | ···················· |
| N | 5170 | hCoV-19/England/QEUH-A5E230/2020\|EPI_ISL_597347\|20 | ···················· | ···················· | ···················· | ···················· | ···················· | ···················· |
| N | 5171 | hCoV-19/England/QEUH-A5E704/2020\|EPI_ISL_597348\|20 | ···················· | ···················· | ···················· | ···················· | ···················· | ···················· |
| N | 5172 | hCoV-19/England/CAMC-A578BD/2020\|EPI_ISL_597349\|20 | ···················· | ···················· | ···················· | ···················· | ···················· | ···················· |
| N | 5173 | hCoV-19/England/CAMC-A57C39/2020\|EPI_ISL_597350\|20 | ···················· | ·A·················· | ···················· | ···················· | ···················· | ···A················ |
| N | 5174 | hCoV-19/England/QEUH-A5E41C/2020\|EPI_ISL_597351\|20 | ···················· | ···················· | ···················· | ···················· | ···················· | ···················· |
| N | 5175 | hCoV-19/England/QEUH-A5DEE2/2020\|EPI_ISL_597352\|20 | ···················· | ·A·················· | ···················· | ···················· | ···················· | ···A················ |
| N | 5176 | hCoV-19/England/QEUH-A5E027/2020\|EPI_ISL_597353\|20 | ···················· | ···················· | ···················· | ···················· | ······T············· | ···················· |
| N | 5177 | hCoV-19/England/CAMC-A572BF/2020\|EPI_ISL_597354\|20 | ···················· | ···················· | ···················· | ···················· | ···················· | ···················· |
| N | 5178 | hCoV-19/England/QEUH-A5E090/2020\|EPI_ISL_597355\|20 | ···················· | ···················· | ···················· | ···················· | ···················· | ···················· |
| N | 5179 | hCoV-19/England/QEUH-A5DE79/2020\|EPI_ISL_597356\|20 | ···················· | ···················· | ···················· | ···················· | ···················· | ···················· |
| N | 5180 | hCoV-19/England/QEUH-A5DF85/2020\|EPI_ISL_597360\|20 | ···················· | ···················· | ···················· | ···················· | ···················· | ···················· |
| N | 5181 | hCoV-19/England/CAMC-A5739E/2020\|EPI_ISL_597361\|20 | ···················· | ···················· | ···················· | ···················· | ···················· | ···················· |
| N | 5182 | hCoV-19/England/QEUH-A5E300/2020\|EPI_ISL_597362\|20 | ···················· | ···················· | ···················· | ···················· | ···················· | ···················· |
| N | 5183 | hCoV-19/England/QEUH-A5DE4C/2020\|EPI_ISL_597379\|20 | ···················· | ···················· | ···················· | ···················· | ···················· | ···················· |
| N | 5184 | hCoV-19/England/CAMC-A574C8/2020\|EPI_ISL_597381\|20 | ···················· | ···················· | ···················· | ···················· | ···················· | ···················· |
| N | 5185 | hCoV-19/England/QEUH-A5E722/2020\|EPI_ISL_597382\|20 | ···················· | ···················· | ···················· | ···················· | ···················· | ···················· |
| N | 5186 | hCoV-19/England/QEUH-A5E90E/2020\|EPI_ISL_597383\|20 | ···················· | ···················· | ···················· | ···················· | ···················· | ···················· |
| N | 5187 | hCoV-19/England/QEUH-A5E35B/2020\|EPI_ISL_597384\|20 | ···················· | ···················· | ···················· | ···················· | ···················· | ···················· |
| N | 5188 | hCoV-19/England/QEUH-A5E494/2020\|EPI_ISL_597385\|20 | ···················· | ···················· | ···················· | ···················· | ···················· | ···················· |
| N | 5189 | hCoV-19/England/QEUH-A5DEF1/2020\|EPI_ISL_597386\|20 | ···················· | ···················· | ···················· | ···················· | ···················· | ···················· |
| N | 5190 | hCoV-19/England/CAMC-A57835/2020\|EPI_ISL_597388\|20 | ···················· | ···················· | ···················· | ···················· | ···················· | ···················· |
| N | 5191 | hCoV-19/England/QEUH-A5DDF4/2020\|EPI_ISL_597389\|20 | ···················· | ···················· | ···················· | ···················· | ···················· | ···················· |
| N | 5192 | hCoV-19/England/CAMC-A574D7/2020\|EPI_ISL_597390\|20 | ···················· | ···················· | ···················· | ···················· | ···················· | ···················· |
| N | 5193 | hCoV-19/England/QEUH-A5E731/2020\|EPI_ISL_597391\|20 | ···················· | ···················· | ···················· | ···················· | ···················· | ···················· |
| N | 5194 | hCoV-19/England/CAMC-A575F2/2020\|EPI_ISL_597392\|20 | ···················· | ···················· | ···················· | ···················· | ···················· | ···················· |
| N | 5195 | hCoV-19/England/QEUH-A5E1D9/2020\|EPI_ISL_597393\|20 | ···················· | ···················· | ···················· | ···················· | ···················· | ···················· |
| N | 5196 | hCoV-19/England/QEUH-A5DF49/2020\|EPI_ISL_597394\|20 | ···················· | ···················· | ···················· | ···················· | ···················· | ···················· |
| N | 5197 | hCoV-19/England/QEUH-A5E4FE/2020\|EPI_ISL_597395\|20 | ···················· | ···················· | ···················· | ···················· | ···················· | ···················· |
| N | 5198 | hCoV-19/England/CAMC-A575A7/2020\|EPI_ISL_597396\|20 | ···················· | ···················· | ···················· | ···················· | ···················· | ···················· |
| N | 5199 | hCoV-19/England/CAMC-A570E2/2020\|EPI_ISL_597397\|20 | ···················· | ···················· | ···················· | ···················· | ···················· | ···················· |
| N | 5200 | hCoV-19/England/CAMC-A56FE6/2020\|EPI_ISL_597398\|20 | ···················· | ···················· | ···················· | ···················· | ···················· | ···················· |
| N | 5201 | hCoV-19/England/CAMC-A57A3F/2020\|EPI_ISL_597399\|20 | ···················· | ···················· | ···················· | ···················· | ···················· | ···················· |
| N | 5202 | hCoV-19/England/MILK-A45D59/2020\|EPI_ISL_598644\|20 | ···················· | ···················· | ···················· | ···················· | ···················· | ···················· |
| N | 5203 | hCoV-19/England/MILK-A461C5/2020\|EPI_ISL_598645\|20 | ···················· | ···················· | ···················· | ···················· | ···················· | ···················· |
| N | 5204 | hCoV-19/England/MILK-A457C4/2020\|EPI_ISL_598646\|20 | ···················· | ···················· | ···················· | ···················· | ···················· | ···················· |
| N | 5205 | hCoV-19/England/MILK-A44F27/2020\|EPI_ISL_598647\|20 | ···················· | ···················· | ···················· | ···················· | ······T············· | ···················· |
| N | 5206 | hCoV-19/England/MILK-A4612F/2020\|EPI_ISL_598648\|20 | ···················· | ···················· | ···················· | ···················· | ···················· | ···················· |
| N | 5207 | hCoV-19/England/MILK-A45F9F/2020\|EPI_ISL_598649\|20 | ···················· | ·A·················· | ···················· | ···················· | ···················· | ···A················ |
| N | 5208 | hCoV-19/England/MILK-A4590A/2020\|EPI_ISL_598651\|20 | ···················· | ···················· | ···················· | ···················· | ···················· | ···················· |
| N | 5209 | hCoV-19/England/MILK-A45BE6/2020\|EPI_ISL_598652\|20 | ···················· | ···················· | ···················· | ···················· | ···················· | ···················· |
| N | 5210 | hCoV-19/England/MILK-A460AA/2020\|EPI_ISL_598653\|20 | ···················· | ···················· | ···················· | ···················· | ···················· | ···················· |
| N | 5211 | hCoV-19/England/MILK-A45A9E/2020\|EPI_ISL_598654\|20 | ···················· | ···················· | ···················· | ···················· | ···················· | ···················· |
| N | 5212 | hCoV-19/England/MILK-A44C6C/2020\|EPI_ISL_598655\|20 | ···················· | ···················· | ···················· | ···················· | ···················· | ···················· |
| N | 5213 | hCoV-19/England/MILK-A4620E/2020\|EPI_ISL_598657\|20 | ···················· | ···················· | ···················· | ···················· | ······T············· | ···················· |
| N | 5214 | hCoV-19/England/MILK-A458D0/2020\|EPI_ISL_598659\|20 | ···················· | ···················· | ···················· | ···················· | ···················· | ···················· |
| N | 5215 | hCoV-19/England/MILK-A44DE1/2020\|EPI_ISL_598660\|20 | ···················· | ···················· | ···················· | ···················· | ···················· | ···················· |
| N | 5216 | hCoV-19/England/MILK-A4670F/2020\|EPI_ISL_598661\|20 | ···················· | ···················· | ···················· | ···················· | ···················· | ···················· |
| N | 5217 | hCoV-19/England/MILK-A463CF/2020\|EPI_ISL_598662\|20 | ···················· | ···················· | ···················· | ···················· | ···················· | ···················· |
| N | 5218 | hCoV-19/England/MILK-A44C02/2020\|EPI_ISL_598663\|20 | ···················· | ···················· | ···················· | ···················· | ···················· | ···················· |
| N | 5219 | hCoV-19/England/MILK-A45876/2020\|EPI_ISL_598664\|20 | ···················· | ···················· | ···················· | ···················· | ···················· | ···················· |
| N | 5220 | hCoV-19/England/MILK-A46480/2020\|EPI_ISL_598665\|20 | ···················· | ···················· | ···················· | ···················· | ···················· | ···················· |
| N | 5221 | hCoV-19/England/MILK-A4630B/2020\|EPI_ISL_598666\|20 | ···················· | ···················· | ···················· | ···················· | ···················· | ···················· |
| N | 5222 | hCoV-19/England/MILK-A46611/2020\|EPI_ISL_598668\|20 | ···················· | ·A·················· | ···················· | ···················· | ···················· | ···A················ |
| N | 5223 | hCoV-19/England/MILK-A45973/2020\|EPI_ISL_598669\|20 | ···················· | ···················· | ···················· | ···················· | ···················· | ···················· |
| N | 5224 | hCoV-19/England/MILK-A456F4/2020\|EPI_ISL_598670\|20 | ···················· | ···················· | ···················· | ···················· | ···················· | ···················· |
| N | 5225 | hCoV-19/England/MILK-A45E29/2020\|EPI_ISL_598671\|20 | ···················· | ···················· | ···················· | ···················· | ···················· | ···················· |

| | | | | | | | | |
|---|---|---|---|---|---|---|---|---|
| N | 5226 | hCoV-19/England/MILK-A46699/2020\|EPI_ISL_598672\|20 | ···················· | ···················· | ···················· | ···················· | ······T············· | ···················· |
| N | 5227 | hCoV-19/England/MILK-A45D3B/2020\|EPI_ISL_598673\|20 | ···················· | ·A·················· | ···················· | ···················· | ···················· | ···A················ |
| N | 5228 | hCoV-19/England/MILK-A45FCC/2020\|EPI_ISL_598674\|20 | ···················· | ···················· | ···················· | ···················· | ···················· | ···················· |
| N | 5229 | hCoV-19/England/MILK-A46620/2020\|EPI_ISL_598675\|20 | ···················· | ···················· | ···················· | ···················· | ···················· | ···················· |
| N | 5230 | hCoV-19/England/MILK-A46875/2020\|EPI_ISL_598676\|20 | ···················· | ···················· | ···················· | ···················· | ···················· | ···················· |
| N | 5231 | hCoV-19/England/MILK-A4583A/2020\|EPI_ISL_598677\|20 | ···················· | ···················· | ···················· | ···················· | ···················· | ···················· |
| N | 5232 | hCoV-19/England/MILK-A44F36/2020\|EPI_ISL_598678\|20 | ···················· | ···················· | ···················· | ···················· | ······T············· | ···················· |
| N | 5233 | hCoV-19/England/MILK-A4674B/2020\|EPI_ISL_598679\|20 | ···················· | ···················· | ···················· | ···················· | ···················· | ···················· |
| N | 5234 | hCoV-19/England/MILK-A44E0C/2020\|EPI_ISL_598680\|20 | ···················· | ···················· | ···················· | ···················· | ···················· | ···················· |
| N | 5235 | hCoV-19/England/MILK-A458A3/2020\|EPI_ISL_598681\|20 | ···················· | ···················· | ···················· | ···················· | ···················· | ···················· |
| N | 5236 | hCoV-19/England/MILK-A45AF8/2020\|EPI_ISL_598682\|20 | ···················· | ···················· | ···················· | ···················· | ···················· | ···················· |
| N | 5237 | hCoV-19/England/MILK-A45DFF/2020\|EPI_ISL_598683\|20 | ···················· | ···················· | ···················· | ···················· | ···················· | ···················· |
| N | 5238 | hCoV-19/England/MILK-A45779/2020\|EPI_ISL_598684\|20 | ···················· | ···················· | ···················· | ···················· | ···················· | ···················· |
| N | 5239 | hCoV-19/England/MILK-A44C99/2020\|EPI_ISL_598685\|20 | ···················· | ···················· | ···················· | ···················· | ···················· | ···················· |
| N | 5240 | hCoV-19/England/MILK-A4573D/2020\|EPI_ISL_598687\|20 | ···················· | ···················· | ···················· | ···················· | ···················· | ···················· |
| N | 5241 | hCoV-19/England/MILK-A45ACB/2020\|EPI_ISL_598692\|20 | ···················· | ···················· | ···················· | ···················· | ···················· | ···················· |
| N | 5242 | hCoV-19/England/MILK-A44BD8/2020\|EPI_ISL_598693\|20 | ···················· | ···················· | ···················· | ···················· | ···················· | ···················· |
| N | 5243 | hCoV-19/England/MILK-A05423/2020\|EPI_ISL_599975\|20 | ···················· | ···················· | ···················· | ···················· | ···················· | ···················· |
| N | 5244 | hCoV-19/England/MILK-A06413/2020\|EPI_ISL_599976\|20 | ···················· | ···················· | ···················· | ···················· | ···················· | ···················· |
| N | 5245 | hCoV-19/England/MILK-A05414/2020\|EPI_ISL_599977\|20 | ···················· | ···················· | ···················· | ···················· | ···················· | ···················· |
| N | 5246 | hCoV-19/England/MILK-A0549C/2020\|EPI_ISL_599978\|20 | ···················· | ···················· | ···················· | ···················· | ···················· | ···················· |
| N | 5247 | hCoV-19/England/MILK-A0564B/2020\|EPI_ISL_599979\|20 | ···················· | ·A·················· | ···················· | ···················· | ···················· | ···A················ |
| N | 5248 | hCoV-19/England/MILK-A061EF/2020\|EPI_ISL_599980\|20 | ···················· | ···················· | ···················· | ···················· | ···················· | ···················· |
| N | 5249 | hCoV-19/England/MILK-A05BC4/2020\|EPI_ISL_599981\|20 | ···················· | ···················· | ···················· | ···················· | ···················· | ···················· |
| N | 5250 | hCoV-19/Wales/MILK-A05836/2020\|EPI_ISL_599982\|2020 | ···················· | ···················· | ···················· | ···················· | ···················· | ···················· |
| N | 5251 | hCoV-19/England/MILK-A056D2/2020\|EPI_ISL_599983\|20 | ···················· | ···················· | ···················· | ···················· | ···················· | ···················· |
| N | 5252 | hCoV-19/England/MILK-A0612B/2020\|EPI_ISL_599984\|20 | ···················· | ···················· | ···················· | ···················· | ···················· | ···················· |
| N | 5253 | hCoV-19/Wales/MILK-A05D91/2020\|EPI_ISL_599985\|2020 | ···················· | ···················· | ···················· | ···················· | ···················· | ···················· |
| N | 5254 | hCoV-19/England/MILK-A06729/2020\|EPI_ISL_599986\|20 | ···················· | ···················· | ···················· | ···················· | ···················· | ···················· |
| N | 5255 | hCoV-19/England/MILK-A06783/2020\|EPI_ISL_599987\|20 | ···················· | ···················· | ···················· | ···················· | ···················· | ···················· |
| N | 5256 | hCoV-19/England/MILK-A06501/2020\|EPI_ISL_599988\|20 | ···················· | ···················· | ···················· | ···················· | ···················· | ···················· |
| N | 5257 | hCoV-19/England/MILK-A057EE/2020\|EPI_ISL_599989\|20 | ···················· | ···················· | ···················· | ···················· | ···················· | ···················· |
| N | 5258 | hCoV-19/England/MILK-A05A7C/2020\|EPI_ISL_599990\|20 | ···················· | ···················· | ···················· | ···················· | ···················· | ···················· |
| N | 5259 | hCoV-19/England/MILK-A06A20/2020\|EPI_ISL_599992\|20 | ···················· | ···················· | ···················· | ···················· | ···················· | ···················· |
| N | 5260 | hCoV-19/England/MILK-A060F1/2020\|EPI_ISL_599993\|20 | ···················· | ···················· | ···················· | ···················· | ···················· | ···················· |
| N | 5261 | hCoV-19/England/MILK-A06307/2020\|EPI_ISL_599994\|20 | ···················· | ···················· | ···················· | ···················· | ···················· | ···················· |
| N | 5262 | hCoV-19/England/MILK-A062A0/2020\|EPI_ISL_599996\|20 | ···················· | ···················· | ···················· | ···················· | ···················· | ···················· |
| N | 5263 | hCoV-19/England/MILK-A06343/2020\|EPI_ISL_599997\|20 | ···················· | ·A·················· | ···················· | ···················· | ···················· | ···A················ |
| N | 5264 | hCoV-19/England/MILK-A05872/2020\|EPI_ISL_599998\|20 | ···················· | ···················· | ···················· | ···················· | ···················· | ···················· |
| N | 5265 | hCoV-19/England/MILK-A05951/2020\|EPI_ISL_599999\|20 | ···················· | ···················· | ···················· | ···················· | ······T············· | ···················· |
| N | 5266 | hCoV-19/Scotland/QEUH-9E4EA5/2020\|EPI_ISL_601000\|2 | ···················· | ···················· | ···················· | ···················· | ···················· | ···················· |
| N | 5267 | hCoV-19/Scotland/QEUH-9E4EE1/2020\|EPI_ISL_601001\|2 | ···················· | ···················· | ···················· | ···················· | ···················· | ···················· |
| N | 5268 | hCoV-19/Scotland/QEUH-9E3F85/2020\|EPI_ISL_601003\|2 | ···················· | ···················· | ···················· | ···················· | ···················· | ···················· |
| N | 5269 | hCoV-19/Scotland/QEUH-9E4D11/2020\|EPI_ISL_601004\|2 | ···················· | ···················· | ···················· | ···················· | ···················· | ···················· |
| N | 5270 | hCoV-19/Scotland/QEUH-9E5439/2020\|EPI_ISL_601005\|2 | ···················· | ·A·················· | ···················· | ···················· | ···················· | ···················· |
| N | 5271 | hCoV-19/Scotland/QEUH-9E5493/2020\|EPI_ISL_601006\|2 | ···················· | ···················· | ···················· | ···················· | ···················· | ···················· |
| N | 5272 | hCoV-19/Scotland/QEUH-9E5581/2020\|EPI_ISL_601007\|2 | ···················· | ···················· | ···················· | ···················· | ···················· | ···················· |
| N | 5273 | hCoV-19/Scotland/QEUH-9E4801/2020\|EPI_ISL_601008\|2 | ···················· | ···················· | ···················· | ···················· | ···················· | ···················· |
| N | 5274 | hCoV-19/Scotland/QEUH-9E5448/2020\|EPI_ISL_601050\|2 | ···················· | ···················· | ···················· | ···················· | ···················· | ···················· |
| N | 5275 | hCoV-19/Scotland/QEUH-9E519C/2020\|EPI_ISL_601051\|2 | ···················· | ···················· | ···················· | ···················· | ···················· | ···················· |
| N | 5276 | hCoV-19/Scotland/QEUH-9E3EB5/2020\|EPI_ISL_601052\|2 | ···················· | ···················· | ···················· | ···················· | ······T············· | ···················· |
| N | 5277 | hCoV-19/Scotland/QEUH-9E55CD/2020\|EPI_ISL_601053\|2 | ···················· | ···················· | ···················· | ···················· | ···················· | ···················· |
| N | 5278 | hCoV-19/Scotland/QEUH-9E5150/2020\|EPI_ISL_601054\|2 | ···················· | ···················· | ···················· | ···················· | ···················· | ···················· |
| N | 5279 | hCoV-19/Scotland/QEUH-9E5132/2020\|EPI_ISL_601055\|2 | ···················· | ···················· | ···················· | ···················· | ···················· | ···················· |
| N | 5280 | hCoV-19/Scotland/QEUH-9E4CE7/2020\|EPI_ISL_601056\|2 | ···················· | ···················· | ···················· | ···················· | ···················· | ···················· |
| N | 5281 | hCoV-19/Scotland/QEUH-9E518D/2020\|EPI_ISL_601057\|2 | ···················· | ···················· | ···················· | ···················· | ···················· | ···················· |
| N | 5282 | hCoV-19/Scotland/QEUH-9E4CD8/2020\|EPI_ISL_601058\|2 | ···················· | ···················· | ···················· | ···················· | ···················· | ···················· |
| N | 5283 | hCoV-19/Scotland/QEUH-9E3F76/2020\|EPI_ISL_601059\|2 | ···················· | ···················· | ···················· | ···················· | ···················· | ···················· |
| N | 5284 | hCoV-19/Scotland/QEUH-9E4740/2020\|EPI_ISL_601060\|2 | ···················· | ···················· | ···················· | ···················· | ···················· | ···················· |
| N | 5285 | hCoV-19/Scotland/QEUH-9E52D5/2020\|EPI_ISL_601062\|2 | ···················· | ···················· | ···················· | ···················· | ···················· | ···················· |
| N | 5286 | hCoV-19/Scotland/QEUH-9E516F/2020\|EPI_ISL_601063\|2 | ···················· | ···················· | ···················· | ···················· | ···················· | ···················· |
| N | 5287 | hCoV-19/Scotland/QEUH-9E41E8/2020\|EPI_ISL_601064\|2 | ···················· | ···················· | ···················· | ···················· | ···················· | ···················· |
| N | 5288 | hCoV-19/Scotland/QEUH-9E44A3/2020\|EPI_ISL_601065\|2 | ···················· | ···················· | ···················· | ···················· | ····T··············· | ···················· |
| N | 5289 | hCoV-19/Scotland/QEUH-9E55DC/2020\|EPI_ISL_601067\|2 | ···················· | ·A·················· | ···················· | ···················· | ···················· | ···················· |
| N | 5290 | hCoV-19/Scotland/QEUH-9E42B8/2020\|EPI_ISL_601068\|2 | ···················· | ···················· | ···················· | ···················· | ···················· | ···················· |
| N | 5291 | hCoV-19/Scotland/QEUH-9E52E4/2020\|EPI_ISL_601069\|2 | ···················· | ···················· | ···················· | ···················· | ···················· | ···················· |
| N | 5292 | hCoV-19/Scotland/QEUH-9E4CC9/2020\|EPI_ISL_601070\|2 | ···················· | ···················· | ···················· | ···················· | ···················· | ···················· |
| N | 5293 | hCoV-19/Scotland/QEUH-9E4F2A/2020\|EPI_ISL_601072\|2 | ···················· | ···················· | ···················· | ···················· | ···················· | ···················· |
| N | 5294 | hCoV-19/Scotland/QEUH-9E485C/2020\|EPI_ISL_601073\|2 | ···················· | ···················· | ···················· | ···················· | ···················· | ···················· |
| N | 5295 | hCoV-19/Scotland/QEUH-9E46BC/2020\|EPI_ISL_601074\|2 | ···················· | ···················· | ···················· | ···················· | ···················· | ···················· |
| N | 5296 | hCoV-19/Scotland/QEUH-9E434C/2020\|EPI_ISL_601075\|2 | ···················· | ···················· | ···················· | ···················· | ···················· | ···················· |
| N | 5297 | hCoV-19/Scotland/QEUH-9E47E6/2020\|EPI_ISL_601076\|2 | ···················· | ···················· | ···················· | ···················· | ···················· | ···················· |
| N | 5298 | hCoV-19/Scotland/QEUH-9E51F6/2020\|EPI_ISL_601077\|2 | ···················· | ···················· | ···················· | ···················· | ···················· | ···················· |
| N | 5299 | hCoV-19/Scotland/QEUH-9E54EE/2020\|EPI_ISL_601078\|2 | ···················· | ···················· | ···················· | ···················· | ···················· | ···················· |
| N | 5300 | hCoV-19/Scotland/QEUH-9E4D3F/2020\|EPI_ISL_601079\|2 | ···················· | ···················· | ···················· | ···················· | ······T············· | ···················· |
| N | 5301 | hCoV-19/Scotland/QEUH-9E48A7/2020\|EPI_ISL_601080\|2 | ···················· | ···················· | ···················· | ···················· | ···················· | ···················· |
| N | 5302 | hCoV-19/Scotland/QEUH-9E4D5D/2020\|EPI_ISL_601081\|2 | ···················· | ···················· | ···················· | ···················· | ···················· | ···················· |
| N | 5303 | hCoV-19/Scotland/QEUH-9E44FE/2020\|EPI_ISL_601082\|2 | ···················· | ···················· | ···················· | ···················· | ···················· | ···················· |
| N | 5304 | hCoV-19/Scotland/QEUH-9E53F0/2020\|EPI_ISL_601083\|2 | ···················· | ···················· | ···················· | ···················· | ···················· | ···················· |
| N | 5305 | hCoV-19/Scotland/QEUH-9E540C/2020\|EPI_ISL_601084\|2 | ···················· | ···················· | ···················· | ···················· | ···················· | ···················· |

```
N 5306 hCoV-19/Scotland/QEUH-9E4081/2020|EPI_ISL_601085|2 .................... .................... .................... .................... .................... ....................
N 5307 hCoV-19/Scotland/QEUH-9E523F/2020|EPI_ISL_601086|2 .................... .................... .................... .................... .................... ....................
N 5308 hCoV-19/Scotland/QEUH-9E527B/2020|EPI_ISL_601087|2 .................... .A.................. .................... .................... .................... ....................
N 5309 hCoV-19/Scotland/QEUH-9E4D6C/2020|EPI_ISL_601088|2 .................... .................... .................... .................... .................... ....................
N 5310 hCoV-19/Scotland/QEUH-9E486B/2020|EPI_ISL_601089|2 .................... .................... .................... .................... .................... ....................
N 5311 hCoV-19/Scotland/QEUH-9E4670/2020|EPI_ISL_601090|2 .................... .................... .................... .................... .................... ....................
N 5312 hCoV-19/Scotland/QEUH-9E4EF0/2020|EPI_ISL_601091|2 .................... .................... .................... .................... .................... ....................
N 5313 hCoV-19/Scotland/QEUH-9E4722/2020|EPI_ISL_601092|2 .................... .................... .................... .................... .................... ....................
N 5314 hCoV-19/Scotland/QEUH-9E417F/2020|EPI_ISL_601093|2 .................... .................... .................... .................... .................... ....................
N 5315 hCoV-19/Scotland/QEUH-9E54DF/2020|EPI_ISL_601094|2 .................... .................... .................... .................... .................... ....................
N 5316 hCoV-19/Scotland/QEUH-9E483E/2020|EPI_ISL_601095|2 .................... .................... .................... .................... .................... ....................
N 5317 hCoV-19/Scotland/QEUH-9E5369/2020|EPI_ISL_601096|2 .................... .................... .................... .................... .................... ....................
N 5318 hCoV-19/Scotland/QEUH-9DC80C/2020|EPI_ISL_601098|2 .................... .................... .................... .................... .................... ....................
N 5319 hCoV-19/USA/FL-BMMG/2020|EPI_ISL_602320|2020-08-31 .................... .................... .................... .................... .................... ....................
N 5320 hCoV-19/USA/PR-B8HT/2020|EPI_ISL_602324|2020-08-12 .................... .................... .................... .................... .................... ....................
N 5321 hCoV-19/Russia/SPE-RII-MH1014S/2020|EPI_ISL_602329 .................... .................... .................... .................... .................... ....................
N 5322 hCoV-19/Russia/SPE-RII-MH1057S/2020|EPI_ISL_602330 .................... .................... .................... .................... .................... ....................
N 5323 hCoV-19/Russia/SVE-RII-MH1442S/2020|EPI_ISL_602331 .................... .................... .................... .................... .................... ....................
N 5324 hCoV-19/Russia/SVE-RII-MH1446S/2020|EPI_ISL_602332 .................... .................... .................... .................... .................... ....................
N 5325 hCoV-19/Russia/SVE-RII-MH1447S/2020|EPI_ISL_602333 .................... .................... .................... .................... .................... ....................
N 5326 hCoV-19/Russia/SVE-RII-MH1448S/2020|EPI_ISL_602334 .................... .................... .................... .................... .................... ....................
N 5327 hCoV-19/Russia/SVE-RII-MH1449S/2020|EPI_ISL_602335 .................... .................... .................... .................... .................... ....................
N 5328 hCoV-19/Russia/SPE-RII-MH1496S/2020|EPI_ISL_602336 .................... .................... .................... .................... .................... ....................
N 5329 hCoV-19/Russia/SPE-RII-MH1529S/2020|EPI_ISL_602337 .................... .................... .................... .................... .................... ....................
N 5330 hCoV-19/Russia/SPE-RII-MH1573S/2020|EPI_ISL_602338 .................... .................... .................... .................... .................... ....................
N 5331 hCoV-19/Russia/SPE-RII-MH1597S/2020|EPI_ISL_602339 .................... .................... .................... .................... .................... ....................
N 5332 hCoV-19/Russia/LEN-RII-MH2021S/2020|EPI_ISL_602380 .................... .................... .................... .................... .................... ....................
N 5333 hCoV-19/Russia/SVE-RII-MH2022S/2020|EPI_ISL_602381 .................... .................... .................... .................... .................... ....................
N 5334 hCoV-19/Russia/SVE-RII-MH2024S/2020|EPI_ISL_602382 .................... .................... .................... .................... .................... ....................
N 5335 hCoV-19/Russia/SVE-RII-MH2025S/2020|EPI_ISL_602383 .................... .................... .................... .................... .................... ....................
N 5336 hCoV-19/Russia/SVE-RII-MH2033S/2020|EPI_ISL_602384 T................... .................... .................... .................... .................... ....................
N 5337 hCoV-19/Russia/SVE-RII-MH2034S/2020|EPI_ISL_602385 .................... .................... .................... .................... .................... ....................
N 5338 hCoV-19/Russia/SVE-RII-MH2038S/2020|EPI_ISL_602386 .................... .................... .................... .................... .................... ....................
N 5339 hCoV-19/Russia/SVE-RII-MH2039S/2020|EPI_ISL_602387 .................... .................... .................... .................... .................... ....................
N 5340 hCoV-19/Russia/SVE-RII-MH2045S/2020|EPI_ISL_602388 .................... .................... .................... .................... .................... ....................
N 5341 hCoV-19/Russia/LEN-RII-MH2050S/2020|EPI_ISL_602389 .................... .................... .................... .................... .................... ....................
N 5342 hCoV-19/Russia/LEN-RII-MH2053S/2020|EPI_ISL_602390 .................... .................... .................... .................... .................... ....................
N 5343 hCoV-19/Russia/MOW-RII-MH2055S/2020|EPI_ISL_602391 .................... .................... .................... .................... .................... ....................
N 5344 hCoV-19/Russia/MOW-RII-MH2056S/2020|EPI_ISL_602392 .................... .................... .................... .................... .................... ....................
N 5345 hCoV-19/Russia/MOW-RII-MH2058S/2020|EPI_ISL_602393 .................... .................... .................... .................... .................... ....................
N 5346 hCoV-19/Russia/MOW-RII-MH2059S/2020|EPI_ISL_602394 .................... .................... .................... .................... .................... ....................
N 5347 hCoV-19/Russia/MOW-RII-MH2060S/2020|EPI_ISL_602395 .................... .................... .................... .................... .................... ....................
N 5348 hCoV-19/Russia/MOW-RII-MH2061S/2020|EPI_ISL_602396 .................... .................... .................... .................... .................... ....................
N 5349 hCoV-19/Russia/MUR-RII-MH2062S/2020|EPI_ISL_602397 .................... .................... .................... .................... .........T.......... ....................
N 5350 hCoV-19/Russia/MUR-RII-MH2063S/2020|EPI_ISL_602398 .................... .................... .................... .................... .................... ....................
N 5351 hCoV-19/Russia/MUR-RII-MH2064S/2020|EPI_ISL_602399 .................... .................... .................... .................... .................... ....................
N 5352 hCoV-19/Switzerland/BL-ETHZ-321409/2020|EPI_ISL_60 .................... .................... .................... .................... .................... ....................
N 5353 hCoV-19/Switzerland/BE-ETHZ-321410/2020|EPI_ISL_60 .................... .................... .................... .................... .................... ....................
N 5354 hCoV-19/Switzerland/ZH-ETHZ-321423/2020|EPI_ISL_60 .................... .A.................. .................... .................... .................... ....................
N 5355 hCoV-19/Switzerland/VD-ETHZ-321424/2020|EPI_ISL_60 .................... .................... .................... .................... .................... ....................
N 5356 hCoV-19/Switzerland/ZH-ETHZ-321425/2020|EPI_ISL_60 .................... .................... .................... .................... ......K............. ....................
N 5357 hCoV-19/Switzerland/VD-ETHZ-321426/2020|EPI_ISL_60 .................... .................... .................... .................... .................... ....................
N 5358 hCoV-19/Switzerland/VS-ETHZ-321427/2020|EPI_ISL_60 .................... .................... .................... .................... .................... ....................
N 5359 hCoV-19/Switzerland/VD-ETHZ-321428/2020|EPI_ISL_60 .................... .................... .................... .................... .................... ....................
N 5360 hCoV-19/Switzerland/VS-ETHZ-321429/2020|EPI_ISL_60 .................... .................... .................... .................... .................... ....................
N 5361 hCoV-19/Switzerland/UR-ETHZ-321442/2020|EPI_ISL_60 .................... .................... .................... .................... .................... ....................
N 5362 hCoV-19/Switzerland/BE-ETHZ-321444/2020|EPI_ISL_60 .................... .................... .................... .................... .................... ....................
N 5363 hCoV-19/Switzerland/GR-ETHZ-321445/2020|EPI_ISL_60 .................... .................... .................... .................... .................... ....................
N 5364 hCoV-19/USA/UT-QDX-2650/2020|EPI_ISL_604980|2020-0 .................... .................... .................... .................... .................... ....................
N 5365 hCoV-19/USA/UT-QDX-2653/2020|EPI_ISL_604981|2020-0 .................... .................... .................... .................... .................... ....................
N 5366 hCoV-19/USA/UT-QDX-2656/2020|EPI_ISL_604982|2020-0 .................... .................... .................... .................... .................... ....................
N 5367 hCoV-19/USA/UT-QDX-2680/2020|EPI_ISL_604990|2020-0 .................... .................... .................... .................... .................... ....................
N 5368 hCoV-19/USA/UT-QDX-2681/2020|EPI_ISL_604991|2020-0 .................... .................... .................... .................... .................... ....................
N 5369 hCoV-19/USA/WI-QDX-2636/2020|EPI_ISL_604992|2020-0 .................... .................... .................... .................... .................... ....................
N 5370 hCoV-19/USA/CA-QDX-2683/2020|EPI_ISL_604993|2020-0 .................... .................... .................... .................... .................... ....................
N 5371 hCoV-19/USA/CA-QDX-2684/2020|EPI_ISL_604994|2020-0 .................... .................... .................... .................... .................... ....................
N 5372 hCoV-19/USA/CA-QDX-2685/2020|EPI_ISL_604995|2020-0 .................... .................... .................... .................... .................... ....................
N 5373 hCoV-19/USA/CA-QDX-2687/2020|EPI_ISL_604996|2020-0 .................... .................... .................... .................... .................... ....................
N 5374 hCoV-19/USA/CA-QDX-2688/2020|EPI_ISL_604997|2020-0 .................... .A.................. .................... .................... .................... ....................
N 5375 hCoV-19/USA/CA-QDX-2691/2020|EPI_ISL_604998|2020-0 .................... .................... .................... .................... .................... ....................
N 5376 hCoV-19/USA/CA-QDX-2692/2020|EPI_ISL_604999|2020-0 .................... .................... .................... .................... .................... ....................
N 5377 hCoV-19/England/CAMC-A57C84/2020|EPI_ISL_597300|20 .................... .................... .................... .................... .................... ....................
N 5378 hCoV-19/England/QEUH-A5E7AA/2020|EPI_ISL_597302|20 .................... .................... .................... .................... .................... ....................
N 5379 hCoV-19/England/QEUH-A5E6BC/2020|EPI_ISL_597303|20 .................... .................... .................... .................... .................... ....................
N 5380 hCoV-19/England/CAMC-A56F9B/2020|EPI_ISL_597304|20 .................... .................... .................... .................... .................... ....................
N 5381 hCoV-19/England/CAMC-A57404/2020|EPI_ISL_597305|20 .................... .................... .................... .................... .................... ....................
N 5382 hCoV-19/England/CAMC-A57237/2020|EPI_ISL_597306|20 .................... .................... .................... .................... .................... ....................
N 5383 hCoV-19/England/CAMC-A570A6/2020|EPI_ISL_597307|20 .................... .................... .................... .................... .................... ....................
N 5384 hCoV-19/England/CAMC-A571EF/2020|EPI_ISL_597308|20 .................... .A.................. .................... .................... .................... ....................
N 5385 hCoV-19/England/CAMC-A57AA8/2020|EPI_ISL_597309|20 .................... .................... .................... .................... .................... ....................
```

| | | | | | | | | |
|---|---|---|---|---|---|---|---|---|
| N | 5386 | hCoV-19/England/CAMC-A57B2D/2020\|EPI_ISL_597310\|20 | ···················· | ···················· | ···················· | ···················· | ···················· | ···················· |
| N | 5387 | hCoV-19/England/QEUH-A5E2B8/2020\|EPI_ISL_597311\|20 | ···················· | ···················· | ···················· | ···················· | ···················· | ···················· |
| N | 5388 | hCoV-19/England/QEUH-A5E5EC/2020\|EPI_ISL_597312\|20 | ···················· | ···················· | ···················· | ···················· | ···················· | ···················· |
| N | 5389 | hCoV-19/England/QEUH-A5E106/2020\|EPI_ISL_597357\|20 | ···················· | ···················· | ···················· | ···················· | ···················· | ···················· |
| N | 5390 | hCoV-19/England/CAMC-A59C28/2020\|EPI_ISL_597358\|20 | ···················· | ···················· | ···················· | ···················· | ···················· | ···················· |
| N | 5391 | hCoV-19/England/QEUH-A5DF94/2020\|EPI_ISL_597359\|20 | ···················· | ···················· | ···················· | ···················· | ···················· | ···················· |
| N | 5392 | hCoV-19/England/QEUH-A5DFA3/2020\|EPI_ISL_597363\|20 | ···················· | ···················· | ···················· | ···················· | ···················· | ···················· |
| N | 5393 | hCoV-19/England/QEUH-A5DD03/2020\|EPI_ISL_597364\|20 | ···················· | ···················· | ···················· | ···················· | ···················· | ···················· |
| N | 5394 | hCoV-19/England/CAMC-A577DE/2020\|EPI_ISL_597366\|20 | ···················· | ···················· | ···················· | ···················· | ···················· | ···················· |
| N | 5395 | hCoV-19/England/CAMC-A57440/2020\|EPI_ISL_597367\|20 | ···················· | ···················· | ···················· | ···················· | ···················· | ···················· |
| N | 5396 | hCoV-19/England/CAMC-A59C46/2020\|EPI_ISL_597368\|20 | ···················· | ·A·················· | ···················· | ···················· | ···················· | ··A················· |
| N | 5397 | hCoV-19/England/QEUH-A5E0DC/2020\|EPI_ISL_597369\|20 | ···················· | ···················· | ···················· | ···················· | ···················· | ···················· |
| N | 5398 | hCoV-19/England/QEUH-A5E142/2020\|EPI_ISL_597370\|20 | ···················· | ···················· | ···················· | ···················· | ···················· | ···················· |
| N | 5399 | hCoV-19/England/CAMC-A59BFE/2020\|EPI_ISL_597371\|20 | ···················· | ···················· | ···················· | ···················· | ···················· | ···················· |
| N | 5400 | hCoV-19/England/CAMC-A576B3/2020\|EPI_ISL_597372\|20 | ···················· | ···················· | ···················· | ···················· | ···················· | ···················· |
| N | 5401 | hCoV-19/England/CAMC-A575E3/2020\|EPI_ISL_597373\|20 | ···················· | ···················· | ···················· | ···················· | ···················· | ···················· |
| N | 5402 | hCoV-19/England/CAMC-A57167/2020\|EPI_ISL_597374\|20 | ···················· | ···················· | ···················· | ···················· | ···················· | ···················· |
| N | 5403 | hCoV-19/England/CAMC-A57510/2020\|EPI_ISL_597375\|20 | ···················· | ···················· | ···················· | ···················· | ···················· | ···················· |
| N | 5404 | hCoV-19/England/QEUH-A5E009/2020\|EPI_ISL_597376\|20 | ···················· | ···················· | ···················· | ···················· | ···················· | ···················· |
| N | 5405 | hCoV-19/England/CAMC-A57B1E/2020\|EPI_ISL_597377\|20 | ···················· | ·A·················· | ···················· | ···················· | ···················· | ···················· |
| N | 5406 | hCoV-19/England/MILK-A44D96/2020\|EPI_ISL_598600\|20 | ···················· | ···················· | ···················· | ···················· | ···················· | ···················· |
| N | 5407 | hCoV-19/England/MILK-A45955/2020\|EPI_ISL_598601\|20 | ···················· | ···················· | ···················· | ···················· | ···················· | ···················· |
| N | 5408 | hCoV-19/England/MILK-A46022/2020\|EPI_ISL_598602\|20 | ···················· | ···················· | ···················· | ···················· | ···················· | ···················· |
| N | 5409 | hCoV-19/England/MILK-A46198/2020\|EPI_ISL_598605\|20 | ···················· | ···················· | ···················· | ···················· | ···················· | ···················· |
| N | 5410 | hCoV-19/England/MILK-A46101/2020\|EPI_ISL_598607\|20 | ···················· | ···················· | ···················· | ···················· | ···················· | ···················· |
| N | 5411 | hCoV-19/England/MILK-A45DD1/2020\|EPI_ISL_598608\|20 | ···················· | ···················· | ···················· | ···················· | ···················· | ···················· |
| N | 5412 | hCoV-19/England/MILK-A44E75/2020\|EPI_ISL_598609\|20 | ···················· | ···················· | ···················· | ···················· | ···················· | ···················· |
| N | 5413 | hCoV-19/England/MILK-A45FBD/2020\|EPI_ISL_598610\|20 | ···················· | ···················· | ···················· | ···················· | ···················· | ···················· |
| N | 5414 | hCoV-19/England/MILK-A45991/2020\|EPI_ISL_598611\|20 | ···················· | ···················· | ···················· | ···················· | ···················· | ···················· |
| N | 5415 | hCoV-19/England/MILK-A44F81/2020\|EPI_ISL_598612\|20 | ···················· | ···················· | ···················· | ···················· | ···················· | ···················· |
| N | 5416 | hCoV-19/England/MILK-A450C9/2020\|EPI_ISL_598613\|20 | ···················· | ···················· | ···················· | ···················· | ···················· | ···················· |
| N | 5417 | hCoV-19/England/MILK-A468A2/2020\|EPI_ISL_598614\|20 | ···················· | ···················· | ···················· | ···················· | ···················· | ···················· |
| N | 5418 | hCoV-19/England/MILK-A462B3/2020\|EPI_ISL_598616\|20 | ···················· | ···················· | ···················· | ···················· | ···················· | ···················· |
| N | 5419 | hCoV-19/England/MILK-A45EDE/2020\|EPI_ISL_598617\|20 | ···················· | ···················· | ···················· | ···················· | ···················· | ···················· |
| N | 5420 | hCoV-19/England/MILK-A464EA/2020\|EPI_ISL_598618\|20 | ···················· | ···················· | ···················· | ···················· | ···················· | ···················· |
| N | 5421 | hCoV-19/England/MILK-A45BB9/2020\|EPI_ISL_598619\|20 | ···················· | ·A·················· | ···················· | ···················· | ···················· | ··A················· |
| N | 5422 | hCoV-19/England/MILK-A45DC2/2020\|EPI_ISL_598621\|20 | ···················· | ·A·················· | ···················· | ···················· | ···················· | ··A················· |
| N | 5423 | hCoV-19/England/MILK-A45005/2020\|EPI_ISL_598622\|20 | ···················· | ···················· | ···················· | ···················· | ···················· | ···················· |
| N | 5424 | hCoV-19/England/MILK-A44FAF/2020\|EPI_ISL_598623\|20 | ···················· | ···················· | ···················· | ···················· | ···················· | ···················· |
| N | 5425 | hCoV-19/England/MILK-A4667B/2020\|EPI_ISL_598624\|20 | ···················· | ···················· | ···················· | ···················· | ···················· | ···················· |
| N | 5426 | hCoV-19/England/MILK-A462A4/2020\|EPI_ISL_598626\|20 | ···················· | ···················· | ···················· | ···················· | ···················· | ···················· |
| N | 5427 | hCoV-19/England/MILK-A4671E/2020\|EPI_ISL_598627\|20 | ···················· | ···················· | ···················· | ···················· | ···················· | ···················· |
| N | 5428 | hCoV-19/England/MILK-A45AE9/2020\|EPI_ISL_598628\|20 | ···················· | ···················· | ···················· | ···················· | ···················· | ···················· |
| N | 5429 | hCoV-19/England/MILK-A466C6/2020\|EPI_ISL_598629\|20 | ···················· | ···················· | ···················· | ···················· | ···················· | ···················· |
| N | 5430 | hCoV-19/England/MILK-A46435/2020\|EPI_ISL_598630\|20 | ···················· | ···················· | ···················· | ···················· | ···················· | ···················· |
| N | 5431 | hCoV-19/England/MILK-A44AAE/2020\|EPI_ISL_598631\|20 | ···················· | ···················· | ···················· | ···················· | ···················· | ···················· |
| N | 5432 | hCoV-19/England/MILK-A4664E/2020\|EPI_ISL_598632\|20 | ···················· | ···················· | ···················· | ···················· | ·····T·············· | ···················· |
| N | 5433 | hCoV-19/England/MILK-A44A71/2020\|EPI_ISL_598634\|20 | ···················· | ···················· | ···················· | ···················· | ···················· | ···················· |
| N | 5434 | hCoV-19/England/MILK-A44C4E/2020\|EPI_ISL_598636\|20 | ···················· | ···················· | ···················· | ···················· | ·····T·············· | ···················· |
| N | 5435 | hCoV-19/England/MILK-A44AF9/2020\|EPI_ISL_598637\|20 | ···················· | ···················· | ···················· | ···················· | ···················· | ···················· |
| N | 5436 | hCoV-19/England/MILK-A46426/2020\|EPI_ISL_598639\|20 | ···················· | ···················· | ···················· | ···················· | ···················· | ···················· |
| N | 5437 | hCoV-19/England/MILK-A44CC6/2020\|EPI_ISL_598640\|20 | ···················· | ···················· | ···················· | ···················· | ···················· | ···················· |
| N | 5438 | hCoV-19/England/MILK-A459A0/2020\|EPI_ISL_598641\|20 | ···················· | ···················· | ···················· | ···················· | ···················· | ···················· |
| N | 5439 | hCoV-19/England/MILK-A45964/2020\|EPI_ISL_598642\|20 | ···················· | ···················· | ···················· | ···················· | ···················· | ···················· |
| N | 5440 | hCoV-19/England/MILK-A45797/2020\|EPI_ISL_598643\|20 | ···················· | ···················· | ···················· | ···················· | ···················· | ···················· |
| N | 5441 | hCoV-19/England/MILK-A4607D/2020\|EPI_ISL_598688\|20 | ···················· | ···················· | ···················· | ···················· | ···················· | ···················· |
| N | 5442 | hCoV-19/England/MILK-A45DE0/2020\|EPI_ISL_598689\|20 | ···················· | ···················· | ···················· | ···················· | ···················· | ···················· |
| N | 5443 | hCoV-19/England/MILK-A467D2/2020\|EPI_ISL_598690\|20 | ···················· | ···················· | ···················· | ···················· | ···················· | ···················· |
| N | 5444 | hCoV-19/England/MILK-A460F5/2020\|EPI_ISL_598694\|20 | ···················· | ···················· | ···················· | ···················· | ···················· | ···················· |
| N | 5445 | hCoV-19/England/MILK-A44DF0/2020\|EPI_ISL_598695\|20 | ···················· | ···················· | ···················· | ···················· | ···················· | ···················· |
| N | 5446 | hCoV-19/England/MILK-A4582B/2020\|EPI_ISL_598696\|20 | ···················· | ···················· | ···················· | ···················· | ···················· | ···················· |
| N | 5447 | hCoV-19/England/MILK-A461B6/2020\|EPI_ISL_598697\|20 | ···················· | ···················· | ···················· | ···················· | ···················· | ···················· |
| N | 5448 | hCoV-19/England/MILK-A45B6E/2020\|EPI_ISL_598698\|20 | ···················· | ···················· | ···················· | ···················· | ···················· | ···················· |
| N | 5449 | hCoV-19/England/MILK-A45F53/2020\|EPI_ISL_598699\|20 | ···················· | ···················· | ···················· | ···················· | ···················· | ···················· |
| N | 5450 | hCoV-19/England/MILK-A0597F/2020\|EPI_ISL_599900\|20 | ···················· | ···················· | ···················· | ···················· | ···················· | ···················· |
| N | 5451 | hCoV-19/England/MILK-A05AC7/2020\|EPI_ISL_599901\|20 | ···················· | ···················· | ···················· | ···················· | ···················· | ···················· |
| N | 5452 | hCoV-19/England/MILK-A06844/2020\|EPI_ISL_599902\|20 | ···················· | ···················· | ···················· | ···················· | ···················· | ···················· |
| N | 5453 | hCoV-19/England/MILK-A05F04/2020\|EPI_ISL_599903\|20 | ···················· | ···················· | ···················· | ···················· | ···················· | ···················· |
| N | 5454 | hCoV-19/England/MILK-A05A4F/2020\|EPI_ISL_599905\|20 | ···················· | ···················· | ···················· | ···················· | ···················· | ···················· |
| N | 5455 | hCoV-19/England/MILK-A05FB9/2020\|EPI_ISL_599906\|20 | ···················· | ·A······A··········· | ···················· | ···················· | ···················· | ···················· |
| N | 5456 | hCoV-19/Wales/MILK-A05DBF/2020\|EPI_ISL_599907\|2020 | ···················· | ···················· | ···················· | ···················· | ···················· | ···················· |
| N | 5457 | hCoV-19/England/MILK-A05D64/2020\|EPI_ISL_599908\|20 | ···················· | ···················· | ···················· | ···················· | ·····T·············· | ···················· |
| N | 5458 | hCoV-19/England/MILK-A05793/2020\|EPI_ISL_599909\|20 | ···················· | ···················· | ···················· | ···················· | ···················· | ···················· |
| N | 5459 | hCoV-19/Wales/MILK-A05B6A/2020\|EPI_ISL_599910\|2020 | ···················· | ·A······A··········· | ···················· | ···················· | ···················· | ···················· |
| N | 5460 | hCoV-19/England/MILK-A0655C/2020\|EPI_ISL_599911\|20 | ···················· | ···················· | ···················· | ···················· | ···················· | ···················· |
| N | 5461 | hCoV-19/England/MILK-A05344/2020\|EPI_ISL_599912\|20 | ···················· | ···················· | ···················· | ···················· | ···················· | ···················· |
| N | 5462 | hCoV-19/England/MILK-A05906/2020\|EPI_ISL_599914\|20 | ···················· | ···················· | ···················· | ···················· | ···················· | ···················· |
| N | 5463 | hCoV-19/England/MILK-A06510/2020\|EPI_ISL_599915\|20 | ···················· | ·A·················· | ···················· | ···················· | ···················· | ··A················· |
| N | 5464 | hCoV-19/England/MILK-A06A8A/2020\|EPI_ISL_599916\|20 | ···················· | ···················· | ···················· | ···················· | ···················· | ···················· |
| N | 5465 | hCoV-19/England/MILK-A05D55/2020\|EPI_ISL_599917\|20 | ···················· | ···················· | ···················· | ···················· | ···················· | ···················· |

| | | | | | | | |
|---|---|---|---|---|---|---|---|
| N | 5466 hCoV-19/England/MILK-A0654D/2020\|EPI_ISL_599918\|20 | .................... | .................... | ..................... | .................... | .................... | .................... |
| N | 5467 hCoV-19/England/MILK-A068DB/2020\|EPI_ISL_599919\|20 | .................... | .................... | ..................... | .................... | .................... | .................... |
| N | 5468 hCoV-19/England/MILK-A061D0/2020\|EPI_ISL_599920\|20 | .................... | .................... | ..................... | .................... | .................... | .................... |
| N | 5469 hCoV-19/England/MILK-A05362/2020\|EPI_ISL_599921\|20 | .................... | .................... | ..................... | .................... | .................... | .................... |
| N | 5470 hCoV-19/England/MILK-A06826/2020\|EPI_ISL_599922\|20 | .................... | .................... | ..................... | .................... | .................... | .................... |
| N | 5471 hCoV-19/England/MILK-A064AA/2020\|EPI_ISL_599923\|20 | .................... | .A.................. | ..................... | .................... | .................... | .................... |
| N | 5472 hCoV-19/England/MILK-A05B00/2020\|EPI_ISL_599924\|20 | .................... | .................... | ..................... | .................... | .................... | .................... |
| N | 5473 hCoV-19/England/MILK-A067ED/2020\|EPI_ISL_599925\|20 | .................... | .................... | ..................... | .................... | .................... | .................... |
| N | 5474 hCoV-19/England/MILK-A06431/2020\|EPI_ISL_599927\|20 | .................... | .................... | ..................... | .................... | .................... | .................... |
| N | 5475 hCoV-19/England/MILK-A05C0D/2020\|EPI_ISL_599928\|20 | .................... | .................... | ..................... | .................... | .................... | .................... |
| N | 5476 hCoV-19/England/MILK-A059CA/2020\|EPI_ISL_599929\|20 | .................... | .................... | ..................... | .................... | .................... | .................... |
| N | 5477 hCoV-19/England/MILK-A05960/2020\|EPI_ISL_599930\|20 | .................... | .A.................. | ..................... | .................... | .................... | ..A................. |
| N | 5478 hCoV-19/England/MILK-A054BA/2020\|EPI_ISL_599931\|20 | .................... | .................... | ..................... | .................... | .................... | .................... |
| N | 5479 hCoV-19/England/MILK-A06176/2020\|EPI_ISL_599932\|20 | .................... | .................... | ..................... | .................... | .................... | .................... |
| N | 5480 hCoV-19/England/MILK-A05E07/2020\|EPI_ISL_599933\|20 | .................... | .A.................. | ..................... | .................... | .................... | ..A................. |
| N | 5481 hCoV-19/England/MILK-A05F5F/2020\|EPI_ISL_599934\|20 | .................... | .................... | ..................... | .................... | .................... | .................... |
| N | 5482 hCoV-19/England/MILK-A05BF1/2020\|EPI_ISL_599935\|20 | .................... | .................... | ..................... | .................... | .................... | .................... |
| N | 5483 hCoV-19/England/MILK-A0652F/2020\|EPI_ISL_599936\|20 | .................... | .................... | ..................... | .................... | .................... | .................... |
| N | 5484 hCoV-19/England/MILK-A05A30/2020\|EPI_ISL_599937\|20 | .................... | .................... | ..................... | .................... | .................... | .................... |
| N | 5485 hCoV-19/England/MILK-A05441/2020\|EPI_ISL_599938\|20 | .................... | .................... | ..................... | .................... | .................... | .................... |
| N | 5486 hCoV-19/England/MILK-A05CEF/2020\|EPI_ISL_599939\|20 | .................... | .................... | ..................... | .................... | .................... | .................... |
| N | 5487 hCoV-19/England/MILK-A06264/2020\|EPI_ISL_599940\|20 | .................... | .................... | ..................... | .................... | .................... | .................... |
| N | 5488 hCoV-19/England/MILK-A06A6C/2020\|EPI_ISL_599941\|20 | .................... | .................... | ..................... | .................... | .................... | .................... |
| N | 5489 hCoV-19/England/MILK-A066C2/2020\|EPI_ISL_599942\|20 | .................... | .................... | ..................... | .................... | .................... | .................... |
| N | 5490 hCoV-19/England/MILK-A06246/2020\|EPI_ISL_599943\|20 | .................... | .................... | ..................... | .................... | .................... | .................... |
| N | 5491 hCoV-19/England/MILK-A05687/2020\|EPI_ISL_599944\|20 | .................... | .................... | ..................... | .................... | .................... | .................... |
| N | 5492 hCoV-19/England/MILK-A06B2D/2020\|EPI_ISL_599945\|20 | .................... | .................... | ..................... | .................... | .................... | .................... |
| N | 5493 hCoV-19/Wales/MILK-A05D73/2020\|EPI_ISL_599946\|2020 | .................... | .................... | ..................... | .................... | .................... | .................... |
| N | 5494 hCoV-19/Wales/MILK-A0562D/2020\|EPI_ISL_599947\|2020 | .................... | .................... | ..................... | .................... | .................... | .................... |
| N | 5495 hCoV-19/England/MILK-A06686/2020\|EPI_ISL_599949\|20 | .................... | .................... | ..................... | .................... | .................... | .................... |
| N | 5496 hCoV-19/England/MILK-A0646E/2020\|EPI_ISL_599951\|20 | .................... | .A.................. | ..................... | .................... | .................... | .................... |
| N | 5497 hCoV-19/Wales/MILK-A05845/2020\|EPI_ISL_599952\|2020 | .................... | .................... | ..................... | .................... | .................... | .................... |
| N | 5498 hCoV-19/England/MILK-A062DD/2020\|EPI_ISL_599953\|20 | .................... | .A.................. | ..................... | .................... | .................... | ..A................. |
| N | 5499 hCoV-19/England/MILK-A05A8B/2020\|EPI_ISL_599954\|20 | .................... | .................... | ..................... | .................... | .................... | .................... |
| N | 5500 hCoV-19/Wales/MILK-A05818/2020\|EPI_ISL_599955\|2020 | .................... | .................... | ..................... | .................... | .................... | .................... |
| N | 5501 hCoV-19/England/MILK-A05E9E/2020\|EPI_ISL_599957\|20 | .................... | .................... | ..................... | .................... | .................... | .................... |
| N | 5502 hCoV-19/England/MILK-A05B1F/2020\|EPI_ISL_599958\|20 | .................... | .................... | ..................... | .................... | .................... | .................... |
| N | 5503 hCoV-19/England/MILK-A05748/2020\|EPI_ISL_599959\|20 | .................... | .A.................. | ..................... | .................... | .................... | ..A................. |
| N | 5504 hCoV-19/England/MILK-A06853/2020\|EPI_ISL_599961\|20 | .................... | .................... | ..................... | .................... | .................... | .................... |
| N | 5505 hCoV-19/England/MILK-A05BE2/2020\|EPI_ISL_599962\|20 | .................... | .................... | ..................... | .................... | .................... | .................... |
| N | 5506 hCoV-19/England/MILK-A05D19/2020\|EPI_ISL_599964\|20 | .................... | .................... | ..................... | .................... | .................... | .................... |
| N | 5507 hCoV-19/England/MILK-A058EB/2020\|EPI_ISL_599965\|20 | .................... | .................... | ..................... | .................... | .................... | .................... |
| N | 5508 hCoV-19/England/MILK-A064C8/2020\|EPI_ISL_599966\|20 | .................... | .................... | ..................... | .................... | .................... | .................... |
| N | 5509 hCoV-19/England/MILK-A053AE/2020\|EPI_ISL_599967\|20 | .................... | .A.................. | ..................... | .................... | .................... | ..A................. |
| N | 5510 hCoV-19/England/MILK-A05CC1/2020\|EPI_ISL_599968\|20 | .................... | .................... | ..................... | .................... | .................... | .................... |
| N | 5511 hCoV-19/England/MILK-A05C67/2020\|EPI_ISL_599969\|20 | .................... | .................... | ..................... | .................... | .................... | .................... |
| N | 5512 hCoV-19/England/MILK-A06149/2020\|EPI_ISL_599970\|20 | .................... | .................... | ..................... | .................... | ......T............. | .................... |
| N | 5513 hCoV-19/Wales/MILK-A065B6/2020\|EPI_ISL_599971\|2020 | .................... | .................... | ..................... | .................... | .................... | .................... |
| N | 5514 hCoV-19/England/MILK-A05C3A/2020\|EPI_ISL_599972\|20 | .................... | .................... | ..................... | .................... | .................... | .................... |
| N | 5515 hCoV-19/Wales/MILK-A05854/2020\|EPI_ISL_599974\|2020 | .................... | .................... | ..................... | .................... | .................... | .................... |
| N | 5516 hCoV-19/USA/TX-DSHS-0928/2020\|EPI_ISL_602226\|2020- | .................... | .................... | ..................... | .................... | .................... | .................... |
| N | 5517 hCoV-19/Switzerland/BL-ETHZ-310480/2020\|EPI_ISL_60 | .................... | .................... | ..................... | .................... | .................... | .................... |
| N | 5518 hCoV-19/Switzerland/AG-ETHZ-310481/2020\|EPI_ISL_60 | .................... | .................... | ..................... | .................... | .................... | .................... |
| N | 5519 hCoV-19/Switzerland/BL-ETHZ-310482/2020\|EPI_ISL_60 | .................... | .................... | ..................... | .................... | ......K............. | .................... |
| N | 5520 hCoV-19/Switzerland/BL-ETHZ-310483/2020\|EPI_ISL_60 | .................... | .................... | ..................... | .................... | .................... | .................... |
| N | 5521 hCoV-19/Switzerland/ZH-ETHZ-310484/2020\|EPI_ISL_60 | .................... | .................... | ..................... | .................... | .................... | .................... |
| N | 5522 hCoV-19/Switzerland/ZH-ETHZ-310485/2020\|EPI_ISL_60 | .................... | .................... | ..................... | .................... | .................... | .................... |
| N | 5523 hCoV-19/Switzerland/BE-ETHZ-310486/2020\|EPI_ISL_60 | .................... | .................... | ..................... | .................... | .................... | .................... |
| N | 5524 hCoV-19/Switzerland/BE-ETHZ-310487/2020\|EPI_ISL_60 | .................... | .................... | ..................... | .................... | .................... | .................... |
| N | 5525 hCoV-19/Switzerland/TG-ETHZ-310488/2020\|EPI_ISL_60 | .................... | .................... | ..................... | .................... | .................... | .................... |
| N | 5526 hCoV-19/Switzerland/SZ-ETHZ-310489/2020\|EPI_ISL_60 | .................... | .................... | ..................... | .................... | .................... | .................... |
| N | 5527 hCoV-19/Switzerland/ZH-ETHZ-310490/2020\|EPI_ISL_60 | .................... | .................... | ..................... | .................... | .................... | .................... |
| N | 5528 hCoV-19/Switzerland/VD-ETHZ-310495/2020\|EPI_ISL_60 | .................... | .................... | ..................... | .................... | .................... | .................... |
| N | 5529 hCoV-19/Switzerland/VD-ETHZ-310496/2020\|EPI_ISL_60 | .................... | .................... | ..................... | .................... | .................... | .................... |
| N | 5530 hCoV-19/Switzerland/VD-ETHZ-310497/2020\|EPI_ISL_60 | .................... | .................... | ..................... | .................... | .................... | .................... |
| N | 5531 hCoV-19/Switzerland/BL-ETHZ-310499/2020\|EPI_ISL_60 | .................... | .................... | ..................... | .................... | .................... | .................... |
| N | 5532 hCoV-19/Switzerland/VD-ETHZ-310500/2020\|EPI_ISL_60 | .................... | .................... | ..................... | .................... | .................... | .................... |
| N | 5533 hCoV-19/Switzerland/BL-ETHZ-310501/2020\|EPI_ISL_60 | .................... | .................... | ..................... | .................... | .................... | .................... |
| N | 5534 hCoV-19/Switzerland/SZ-ETHZ-321350/2020\|EPI_ISL_60 | .................... | .................... | ..................... | .................... | .................... | .................... |
| N | 5535 hCoV-19/Switzerland/SZ-ETHZ-321351/2020\|EPI_ISL_60 | .................... | .................... | ..................... | .................... | .................... | .................... |
| N | 5536 hCoV-19/Switzerland/BE-ETHZ-321352/2020\|EPI_ISL_60 | .................... | .................... | ..................... | .................... | .................... | .................... |
| N | 5537 hCoV-19/Switzerland/SZ-ETHZ-321353/2020\|EPI_ISL_60 | .................... | .................... | ..................... | .................... | .................... | .................... |
| N | 5538 hCoV-19/Switzerland/SZ-ETHZ-321354/2020\|EPI_ISL_60 | .................... | .................... | ..................... | .................... | .................... | .................... |
| N | 5539 hCoV-19/Switzerland/AG-ETHZ-321356/2020\|EPI_ISL_60 | .................... | .................... | ..................... | .................... | .................... | .................... |
| N | 5540 hCoV-19/Switzerland/VD-ETHZ-321357/2020\|EPI_ISL_60 | .................... | .................... | ..................... | .................... | .................... | .................... |
| N | 5541 hCoV-19/Switzerland/VD-ETHZ-321358/2020\|EPI_ISL_60 | .................... | .................... | ..................... | .................... | .................... | .................... |
| N | 5542 hCoV-19/Switzerland/VD-ETHZ-321360/2020\|EPI_ISL_60 | .................... | .................... | ..................... | .................... | ......K............. | .................... |
| N | 5543 hCoV-19/Switzerland/VD-ETHZ-321361/2020\|EPI_ISL_60 | .................... | .................... | ..................... | .................... | .................... | .................... |
| N | 5544 hCoV-19/Switzerland/VD-ETHZ-321362/2020\|EPI_ISL_60 | .................... | .................... | ..................... | .................... | .................... | .................... |
| N | 5545 hCoV-19/Switzerland/VD-ETHZ-321363/2020\|EPI_ISL_60 | .................... | .................... | ..................... | .................... | .................... | .................... |

| | | | | | | | |
|---|---|---|---|---|---|---|---|
| N | 5546 hCoV-19/Switzerland/VD-ETHZ-321364/2020\|EPI_ISL_60 | ···················· | ···················· | ···················· | ···················· | ···················· | ···················· |
| N | 5547 hCoV-19/Switzerland/VD-ETHZ-321365/2020\|EPI_ISL_60 | ···················· | ···················· | ···················· | ···················· | ···················· | ···················· |
| N | 5548 hCoV-19/Switzerland/VD-ETHZ-321366/2020\|EPI_ISL_60 | ···················· | ···················· | ···················· | ···················· | ···················· | ···················· |
| N | 5549 hCoV-19/Switzerland/VD-ETHZ-321367/2020\|EPI_ISL_60 | ···················· | ···················· | ···················· | ···················· | ·····K·············· | ···················· |
| N | 5550 hCoV-19/Switzerland/VD-ETHZ-321368/2020\|EPI_ISL_60 | ···················· | ···················· | ···················· | ···················· | ···················· | ···················· |
| N | 5551 hCoV-19/Switzerland/VD-ETHZ-321370/2020\|EPI_ISL_60 | ···················· | ···················· | ···················· | ···················· | ···················· | ···················· |
| N | 5552 hCoV-19/Switzerland/ZH-ETHZ-321371/2020\|EPI_ISL_60 | ···················· | ···················· | ···················· | ···················· | ·····K·············· | ···················· |
| N | 5553 hCoV-19/Switzerland/BS-ETHZ-321374/2020\|EPI_ISL_60 | ···················· | ·A·················· | ···················· | ···················· | ···················· | ···················· |
| N | 5554 hCoV-19/Switzerland/SZ-ETHZ-321376/2020\|EPI_ISL_60 | ···················· | ···················· | ···················· | ···················· | ···················· | ···················· |
| N | 5555 hCoV-19/Switzerland/TI-ETHZ-321378/2020\|EPI_ISL_60 | ···················· | ···················· | ···················· | ···················· | ·····K·············· | ···················· |
| N | 5556 hCoV-19/Switzerland/UR-ETHZ-321379/2020\|EPI_ISL_60 | ···················· | ···················· | ···················· | ···················· | ···················· | ···················· |
| N | 5557 hCoV-19/Switzerland/UR-ETHZ-321380/2020\|EPI_ISL_60 | ···················· | ···················· | ···················· | ···················· | ···················· | ···················· |
| N | 5558 hCoV-19/Switzerland/VD-ETHZ-321382/2020\|EPI_ISL_60 | ···················· | ···················· | ···················· | ···················· | ···················· | ···················· |
| N | 5559 hCoV-19/Switzerland/VD-ETHZ-321383/2020\|EPI_ISL_60 | ···················· | ···················· | ···················· | ···················· | ···················· | ···················· |
| N | 5560 hCoV-19/USA/CA-QDX-2585/2020\|EPI_ISL_604880\|2020-0 | ···················· | ···················· | ···················· | ···················· | ···················· | ···················· |
| N | 5561 hCoV-19/USA/CA-QDX-2586/2020\|EPI_ISL_604881\|2020-0 | ···················· | ···················· | ···················· | ···················· | ···················· | ···················· |
| N | 5562 hCoV-19/USA/CA-QDX-2591/2020\|EPI_ISL_604882\|2020-0 | ···················· | ···················· | ···················· | ···················· | ···················· | ···················· |
| N | 5563 hCoV-19/USA/CA-QDX-2592/2020\|EPI_ISL_604883\|2020-0 | ···················· | ···················· | ···················· | ···················· | ···················· | ···················· |
| N | 5564 hCoV-19/USA/CA-QDX-2593/2020\|EPI_ISL_604884\|2020-0 | ···················· | ···················· | ···················· | ···················· | ···················· | ···················· |
| N | 5565 hCoV-19/USA/CA-QDX-2594/2020\|EPI_ISL_604885\|2020-0 | ···················· | ···················· | ···················· | ···················· | ···················· | ···················· |
| N | 5566 hCoV-19/USA/CA-QDX-2597/2020\|EPI_ISL_604886\|2020-0 | ···················· | ···················· | ···················· | ···················· | ···················· | ···················· |
| N | 5567 hCoV-19/USA/CA-QDX-2600/2020\|EPI_ISL_604887\|2020-0 | ···················· | ···················· | ···················· | ···················· | ···················· | ···················· |
| N | 5568 hCoV-19/USA/CA-QDX-2601/2020\|EPI_ISL_604888\|2020-0 | ···················· | ···················· | ···················· | ···················· | ···················· | ···················· |
| N | 5569 hCoV-19/USA/CA-QDX-2602/2020\|EPI_ISL_604889\|2020-0 | ···················· | ···················· | ···················· | ···················· | ···················· | ···················· |
| N | 5570 hCoV-19/USA/CA-QDX-2604/2020\|EPI_ISL_604891\|2020-0 | ···················· | ···················· | ···················· | ···················· | ···················· | ···················· |
| N | 5571 hCoV-19/USA/CA-QDX-2608/2020\|EPI_ISL_604892\|2020-0 | ···················· | ···················· | ···················· | ···················· | ···················· | ···················· |
| N | 5572 hCoV-19/USA/CA-QDX-2615/2020\|EPI_ISL_604895\|2020-0 | ···················· | ···················· | ···················· | ···················· | ···················· | ···················· |
| N | 5573 hCoV-19/USA/CA-QDX-2620/2020\|EPI_ISL_604896\|2020-0 | ···················· | ···················· | ···················· | ···················· | ···················· | ···················· |
| N | 5574 hCoV-19/USA/CA-QDX-2623/2020\|EPI_ISL_604897\|2020-0 | ···················· | ···················· | ···················· | ···················· | ···················· | ···················· |
| N | 5575 hCoV-19/USA/CA-QDX-2625/2020\|EPI_ISL_604898\|2020-0 | ···················· | ···················· | ···················· | ···················· | ···················· | ···················· |
| N | 5576 hCoV-19/USA/CA-QDX-2629/2020\|EPI_ISL_604899\|2020-0 | ···················· | ···················· | ···················· | ···················· | ···················· | ···················· |
| N | 5577 hCoV-19/USA/CA-QDX-2630/2020\|EPI_ISL_604900\|2020-0 | ···················· | ···················· | ···················· | ···················· | ···················· | ···················· |
| N | 5578 hCoV-19/USA/CA-QDX-2633/2020\|EPI_ISL_604901\|2020-0 | ···················· | ···················· | ···················· | ···················· | ···················· | ···················· |
| N | 5579 hCoV-19/USA/CA-QDX-2637/2020\|EPI_ISL_604902\|2020-0 | ···················· | ···················· | ···················· | ···················· | ···················· | ···················· |
| N | 5580 hCoV-19/USA/CA-QDX-2639/2020\|EPI_ISL_604903\|2020-0 | ···················· | ···················· | ···················· | ···················· | ···················· | ···················· |
| N | 5581 hCoV-19/USA/CA-QDX-2640/2020\|EPI_ISL_604904\|2020-0 | ···················· | ···················· | ···················· | ···················· | ···················· | ···················· |
| N | 5582 hCoV-19/USA/CA-QDX-2641/2020\|EPI_ISL_604905\|2020-0 | ···················· | ···················· | ···················· | ···················· | ···················· | ···················· |
| N | 5583 hCoV-19/USA/CA-QDX-2642/2020\|EPI_ISL_604906\|2020-0 | ···················· | ···················· | ···················· | ···················· | ···················· | ···················· |
| N | 5584 hCoV-19/USA/CA-QDX-2645/2020\|EPI_ISL_604907\|2020-0 | ···················· | ···················· | ···················· | ···················· | ···················· | ···················· |
| N | 5585 hCoV-19/USA/CA-QDX-2646/2020\|EPI_ISL_604908\|2020-0 | ···················· | ···················· | ···················· | ···················· | ···················· | ···················· |
| N | 5586 hCoV-19/USA/CA-QDX-2647/2020\|EPI_ISL_604909\|2020-0 | ···················· | ···················· | ···················· | ···················· | ···················· | ···················· |
| N | 5587 hCoV-19/USA/CA-QDX-2654/2020\|EPI_ISL_604910\|2020-0 | ···················· | ···················· | ···················· | ···················· | ···················· | ···················· |
| N | 5588 hCoV-19/USA/CA-QDX-2658/2020\|EPI_ISL_604911\|2020-0 | ···················· | ···················· | ···················· | ···················· | ···················· | ···················· |
| N | 5589 hCoV-19/USA/CA-QDX-2660/2020\|EPI_ISL_604912\|2020-0 | ···················· | ···················· | ···················· | ···················· | ···················· | ···················· |
| N | 5590 hCoV-19/USA/CA-QDX-2665/2020\|EPI_ISL_604913\|2020-0 | ···················· | ···················· | ···················· | ···················· | ···················· | ···················· |
| N | 5591 hCoV-19/USA/CA-QDX-2674/2020\|EPI_ISL_604914\|2020-0 | ···················· | ···················· | ···················· | ···················· | ···················· | ···················· |
| N | 5592 hCoV-19/USA/CA-QDX-2675/2020\|EPI_ISL_604915\|2020-0 | ···················· | ···················· | ···················· | ···················· | ···················· | ···················· |
| N | 5593 hCoV-19/USA/CA-QDX-2676/2020\|EPI_ISL_604916\|2020-0 | ···················· | ···················· | ···················· | ···················· | ···················· | ···················· |
| N | 5594 hCoV-19/USA/CA-QDX-2677/2020\|EPI_ISL_604917\|2020-0 | ···················· | ···················· | ···················· | ···················· | ···················· | ···················· |
| N | 5595 hCoV-19/USA/CA-QDX-2678/2020\|EPI_ISL_604918\|2020-0 | ···················· | ···················· | ···················· | ···················· | ···················· | ···················· |
| N | 5596 hCoV-19/USA/CT-QDX-2579/2020\|EPI_ISL_604919\|2020-0 | ···················· | ···················· | ···················· | ···················· | ···················· | ···················· |
| N | 5597 hCoV-19/USA/CT-QDX-2666/2020\|EPI_ISL_604920\|2020-0 | ···················· | ···················· | ···················· | ···················· | ···················· | ···················· |
| N | 5598 hCoV-19/USA/FL-QDX-2564/2020\|EPI_ISL_604922\|2020-0 | ···················· | ···················· | ···················· | ···················· | ···················· | ···················· |
| N | 5599 hCoV-19/USA/FL-QDX-2577/2020\|EPI_ISL_604923\|2020-0 | ···················· | ···················· | ···················· | ···················· | ···················· | ···················· |
| N | 5600 hCoV-19/USA/FL-QDX-2588/2020\|EPI_ISL_604924\|2020-0 | ···················· | ···················· | ···················· | ···················· | ···················· | ···················· |
| N | 5601 hCoV-19/USA/FL-QDX-2590/2020\|EPI_ISL_604925\|2020-0 | ···················· | ···················· | ···················· | ···················· | ···················· | ···················· |
| N | 5602 hCoV-19/USA/FL-QDX-2617/2020\|EPI_ISL_604926\|2020-0 | ···················· | ···················· | ···················· | ···················· | ···················· | ···················· |
| N | 5603 hCoV-19/USA/FL-QDX-2618/2020\|EPI_ISL_604927\|2020-0 | ···················· | ···················· | ···················· | ···················· | ···················· | ···················· |
| N | 5604 hCoV-19/USA/FL-QDX-2648/2020\|EPI_ISL_604928\|2020-0 | ···················· | ···················· | ···················· | ···················· | ···················· | ···················· |
| N | 5605 hCoV-19/USA/FL-QDX-2659/2020\|EPI_ISL_604929\|2020-0 | ···················· | ···················· | ···················· | ···················· | ···················· | ···················· |
| N | 5606 hCoV-19/USA/FL-QDX-2670/2020\|EPI_ISL_604930\|2020-0 | ···················· | ···················· | ···················· | ···················· | ···················· | ···················· |
| N | 5607 hCoV-19/USA/GA-QDX-2571/2020\|EPI_ISL_604931\|2020-0 | ···················· | ···················· | ···················· | ···················· | ···················· | ···················· |
| N | 5608 hCoV-19/USA/IL-QDX-2568/2020\|EPI_ISL_604932\|2020-0 | ···················· | ···················· | ···················· | ···················· | ···················· | ···················· |
| N | 5609 hCoV-19/USA/IL-QDX-2622/2020\|EPI_ISL_604933\|2020-0 | ···················· | ···················· | ···················· | ···················· | ···················· | ···················· |
| N | 5610 hCoV-19/USA/IN-QDX-2619/2020\|EPI_ISL_604935\|2020-0 | ···················· | ···················· | ···················· | ···················· | ···················· | ···················· |
| N | 5611 hCoV-19/USA/LA-QDX-2624/2020\|EPI_ISL_604936\|2020-0 | ···················· | ···················· | ···················· | ···················· | ···················· | ···················· |
| N | 5612 hCoV-19/USA/MN-QDX-2565/2020\|EPI_ISL_604937\|2020-0 | ···················· | ···················· | ···················· | ···················· | ···················· | ···················· |
| N | 5613 hCoV-19/USA/MN-QDX-2611/2020\|EPI_ISL_604938\|2020-0 | ···················· | ···················· | ···················· | ···················· | ···················· | ···················· |
| N | 5614 hCoV-19/USA/MO-QDX-2566/2020\|EPI_ISL_604939\|2020-0 | ···················· | ···················· | ···················· | ···················· | ···················· | ···················· |
| N | 5615 hCoV-19/USA/MO-QDX-2567/2020\|EPI_ISL_604940\|2020-0 | ···················· | ···················· | ···················· | ···················· | ···················· | ···················· |
| N | 5616 hCoV-19/USA/MO-QDX-2573/2020\|EPI_ISL_604941\|2020-0 | ···················· | ···················· | ···················· | ···················· | ···················· | ···················· |
| N | 5617 hCoV-19/USA/MO-QDX-2574/2020\|EPI_ISL_604942\|2020-0 | ···················· | ···················· | ···················· | ···················· | ···················· | ···················· |
| N | 5618 hCoV-19/USA/MO-QDX-2582/2020\|EPI_ISL_604943\|2020-0 | ···················· | ···················· | ···················· | ···················· | ···················· | ···················· |
| N | 5619 hCoV-19/USA/MO-QDX-2587/2020\|EPI_ISL_604944\|2020-0 | ···················· | ···················· | ···················· | ···················· | ···················· | ···················· |
| N | 5620 hCoV-19/USA/MO-QDX-2595/2020\|EPI_ISL_604945\|2020-0 | ···················· | ···················· | ···················· | ···················· | ···················· | ···················· |
| N | 5621 hCoV-19/USA/MO-QDX-2612/2020\|EPI_ISL_604946\|2020-0 | ···················· | ···················· | ···················· | ···················· | ···················· | ···················· |
| N | 5622 hCoV-19/USA/MO-QDX-2613/2020\|EPI_ISL_604947\|2020-0 | ···················· | ···················· | ···················· | ···················· | ···················· | ···················· |
| N | 5623 hCoV-19/USA/MO-QDX-2616/2020\|EPI_ISL_604948\|2020-0 | ···················· | ···················· | ···················· | ···················· | ···················· | ···················· |
| N | 5624 hCoV-19/USA/MO-QDX-2679/2020\|EPI_ISL_604949\|2020-0 | ···················· | ···················· | ···················· | ···················· | ···················· | ···················· |
| N | 5625 hCoV-19/USA/NV-QDX-2627/2020\|EPI_ISL_604950\|2020-0 | ···················· | ···················· | ···················· | ···················· | ···················· | ···················· |

| | | | | | | | | |
|---|---|---|---|---|---|---|---|---|
| N | 5626 | hCoV-19/USA/NV-QDX-2631/2020\|EPI_ISL_604951\|2020-0 | ···················· | ···················· | ····················· | ···················· | ···················· | ···················· |
| N | 5627 | hCoV-19/USA/NV-QDX-2635/2020\|EPI_ISL_604952\|2020-0 | ···················· | ···················· | ····················· | ···················· | ···················· | ···················· |
| N | 5628 | hCoV-19/USA/NV-QDX-2652/2020\|EPI_ISL_604953\|2020-0 | ···················· | ···················· | ····················· | ···················· | ···················· | ···················· |
| N | 5629 | hCoV-19/USA/NV-QDX-2662/2020\|EPI_ISL_604954\|2020-0 | ···················· | ···················· | ····················· | ···················· | ···················· | ···················· |
| N | 5630 | hCoV-19/USA/NY-QDX-2562/2020\|EPI_ISL_604955\|2020-0 | ···················· | ···················· | ····················· | ···················· | ···················· | ···················· |
| N | 5631 | hCoV-19/USA/NY-QDX-2651/2020\|EPI_ISL_604956\|2020-0 | ···················· | ···················· | ····················· | ···················· | ···················· | ···················· |
| N | 5632 | hCoV-19/USA/NY-QDX-2661/2020\|EPI_ISL_604957\|2020-0 | ···················· | ···················· | ····················· | ···················· | ···················· | ···················· |
| N | 5633 | hCoV-19/England/CAMC-A59745/2020\|EPI_ISL_597160\|20 | ···················· | ···················· | ····················· | ···················· | ···················· | ···················· |
| N | 5634 | hCoV-19/England/CAMC-A594B7/2020\|EPI_ISL_597162\|20 | ···················· | ···················· | ····················· | ···················· | ···················· | ···················· |
| N | 5635 | hCoV-19/England/CAMC-A58AE3/2020\|EPI_ISL_597163\|20 | ···················· | ···················· | ····················· | ···················· | ···················· | ···················· |
| N | 5636 | hCoV-19/England/CAMC-A5936F/2020\|EPI_ISL_597164\|20 | ···················· | ·A·················· | ····················· | ···················· | ···················· | ···A················ |
| N | 5637 | hCoV-19/England/CAMC-A593C9/2020\|EPI_ISL_597165\|20 | ···················· | ···················· | ····················· | ···················· | ···················· | ···················· |
| N | 5638 | hCoV-19/England/CAMC-A58834/2020\|EPI_ISL_597166\|20 | ···················· | ···················· | ····················· | ···················· | ···················· | ···················· |
| N | 5639 | hCoV-19/England/CAMC-A59587/2020\|EPI_ISL_597167\|20 | ···················· | ···················· | ····················· | ···················· | ···················· | ···················· |
| N | 5640 | hCoV-19/England/CAMC-A57EBB/2020\|EPI_ISL_597168\|20 | ···················· | ···················· | ····················· | ···················· | ···················· | ···················· |
| N | 5641 | hCoV-19/England/CAMC-A5846D/2020\|EPI_ISL_597169\|20 | ···················· | ···················· | ····················· | ···················· | ···················· | ···················· |
| N | 5642 | hCoV-19/England/CAMC-A58816/2020\|EPI_ISL_597170\|20 | ···················· | ···················· | ····················· | ···················· | ···················· | ···················· |
| N | 5643 | hCoV-19/England/CAMC-A58DDB/2020\|EPI_ISL_597171\|20 | ···················· | ···················· | ····················· | ···················· | ···················· | ···················· |
| N | 5644 | hCoV-19/England/CAMC-A59763/2020\|EPI_ISL_597172\|20 | ···················· | ···················· | ····················· | ···················· | ···················· | ···················· |
| N | 5645 | hCoV-19/England/CAMC-A57ED9/2020\|EPI_ISL_597173\|20 | ···················· | ···················· | ····················· | ···················· | ···················· | ···················· |
| N | 5646 | hCoV-19/England/CAMC-A5938D/2020\|EPI_ISL_597174\|20 | ···················· | ···················· | ····················· | ···················· | ···················· | ···················· |
| N | 5647 | hCoV-19/England/CAMC-A594F3/2020\|EPI_ISL_597175\|20 | ···················· | ···················· | ····················· | ···················· | ···················· | ···················· |
| N | 5648 | hCoV-19/England/CAMC-A58F7B/2020\|EPI_ISL_597176\|20 | ···················· | ···················· | ····················· | ···················· | ···················· | ···················· |
| N | 5649 | hCoV-19/England/CAMC-A584C7/2020\|EPI_ISL_597177\|20 | ···················· | ···················· | ····················· | ···················· | ···················· | ···················· |
| N | 5650 | hCoV-19/England/CAMC-A57F21/2020\|EPI_ISL_597178\|20 | ···················· | ···················· | ····················· | ···················· | ···················· | ···················· |
| N | 5651 | hCoV-19/England/CAMC-A59675/2020\|EPI_ISL_597179\|20 | ···················· | ···················· | ····················· | ···················· | ···················· | ···················· |
| N | 5652 | hCoV-19/England/CAMC-A57F9A/2020\|EPI_ISL_597180\|20 | ···················· | ···················· | ····················· | ···················· | ···················· | ···················· |
| N | 5653 | hCoV-19/England/CAMC-A58E6F/2020\|EPI_ISL_597181\|20 | ···················· | ···················· | ····················· | ···················· | ···················· | ···················· |
| N | 5654 | hCoV-19/England/CAMC-A58CDE/2020\|EPI_ISL_597182\|20 | ···················· | ···················· | ····················· | ···················· | ···················· | ···················· |
| N | 5655 | hCoV-19/England/CAMC-A585C4/2020\|EPI_ISL_597183\|20 | ···················· | ···················· | ····················· | ···················· | ···················· | ···················· |
| N | 5656 | hCoV-19/England/CAMC-A593F6/2020\|EPI_ISL_597184\|20 | ···················· | ···················· | ····················· | ···················· | ···················· | ···················· |
| N | 5657 | hCoV-19/England/CAMC-A58184/2020\|EPI_ISL_597185\|20 | ···················· | ···················· | ····················· | ···················· | ···················· | ···················· |
| N | 5658 | hCoV-19/England/CAMC-A58315/2020\|EPI_ISL_597186\|20 | ···················· | ···················· | ····················· | ···················· | ···················· | ···················· |
| N | 5659 | hCoV-19/England/CAMC-A5837F/2020\|EPI_ISL_597187\|20 | ···················· | ···················· | ····················· | ···················· | ···················· | ···················· |
| N | 5660 | hCoV-19/England/CAMC-A594E4/2020\|EPI_ISL_597188\|20 | ···················· | ···················· | ····················· | ···················· | ···················· | ···················· |
| N | 5661 | hCoV-19/England/CAMC-A58E23/2020\|EPI_ISL_597190\|20 | ···················· | ···················· | ····················· | ···················· | ···················· | ···················· |
| N | 5662 | hCoV-19/England/CAMC-A58AA7/2020\|EPI_ISL_597192\|20 | ···················· | ···················· | ····················· | ···················· | ···················· | ···················· |
| N | 5663 | hCoV-19/England/CAMC-A5854C/2020\|EPI_ISL_597193\|20 | ···················· | ···················· | ····················· | ···················· | ···················· | ···················· |
| N | 5664 | hCoV-19/England/CAMC-A594A8/2020\|EPI_ISL_597195\|20 | ···················· | ···················· | ····················· | ···················· | ···················· | ···················· |
| N | 5665 | hCoV-19/England/CAMC-A587A0/2020\|EPI_ISL_597196\|20 | ···················· | ···················· | ····················· | ···················· | ···················· | ···················· |
| N | 5666 | hCoV-19/England/CAMC-A58EBA/2020\|EPI_ISL_597197\|20 | ···················· | ···················· | ····················· | ···················· | ···················· | ···················· |
| N | 5667 | hCoV-19/England/CAMC-A58DAE/2020\|EPI_ISL_597199\|20 | ···················· | ···················· | ····················· | ···················· | ···················· | ···················· |
| N | 5668 | hCoV-19/England/CAMC-A58078/2020\|EPI_ISL_597214\|20 | ···················· | ···················· | ····················· | ···················· | ···················· | ···················· |
| N | 5669 | hCoV-19/England/CAMC-A57D63/2020\|EPI_ISL_597215\|20 | ···················· | ·A·················· | ····················· | ···················· | ···················· | ···A················ |
| N | 5670 | hCoV-19/England/CAMC-A59411/2020\|EPI_ISL_597216\|20 | ···················· | ···················· | ····················· | ···················· | ···················· | ···················· |
| N | 5671 | hCoV-19/England/CAMC-A58272/2020\|EPI_ISL_597217\|20 | ···················· | ·A·······A·········· | ····················· | ···················· | ···················· | ···················· |
| N | 5672 | hCoV-19/England/CAMC-A58B1D/2020\|EPI_ISL_597218\|20 | ···················· | ···················· | ····················· | ···················· | ···················· | ···················· |
| N | 5673 | hCoV-19/England/CAMC-A592DB/2020\|EPI_ISL_597219\|20 | ···················· | ···················· | ····················· | ···················· | ···················· | ···················· |
| N | 5674 | hCoV-19/England/CAMC-A58148/2020\|EPI_ISL_597220\|20 | ···················· | ···················· | ····················· | ···················· | ···················· | ···················· |
| N | 5675 | hCoV-19/England/CAMC-A58DCC/2020\|EPI_ISL_597221\|20 | ···················· | ···················· | ····················· | ···················· | ···················· | ···················· |
| N | 5676 | hCoV-19/England/CAMC-A598CA/2020\|EPI_ISL_597222\|20 | ···················· | ···················· | ····················· | ···················· | ···················· | ···················· |
| N | 5677 | hCoV-19/England/CAMC-A58755/2020\|EPI_ISL_597224\|20 | ···················· | ···················· | ····················· | ···················· | ···················· | ···················· |
| N | 5678 | hCoV-19/England/CAMC-A58782/2020\|EPI_ISL_597230\|20 | ···················· | ···················· | ····················· | ···················· | ···················· | ···················· |
| N | 5679 | hCoV-19/England/CAMC-A58843/2020\|EPI_ISL_597232\|20 | ···················· | ···················· | ····················· | ···················· | ······T············· | ···················· |
| N | 5680 | hCoV-19/England/CAMC-A57176/2020\|EPI_ISL_597233\|20 | ············Y······· | ···················· | ····················· | ···················· | ···················· | ···················· |
| N | 5681 | hCoV-19/England/QEUH-A5E582/2020\|EPI_ISL_597234\|20 | ···················· | ···················· | ····················· | ···················· | ······T············· | ···················· |
| N | 5682 | hCoV-19/England/MILK-A47908/2020\|EPI_ISL_598490\|20 | ···················· | ···················· | ····················· | ···················· | ···················· | ···················· |
| N | 5683 | hCoV-19/England/MILK-A4805D/2020\|EPI_ISL_598491\|20 | ···················· | ···················· | ····················· | ···················· | ···················· | ···················· |
| N | 5684 | hCoV-19/England/MILK-A46A60/2020\|EPI_ISL_598492\|20 | ···················· | ···················· | ····················· | ···················· | ···················· | ···················· |
| N | 5685 | hCoV-19/England/MILK-A471C4/2020\|EPI_ISL_598493\|20 | ···················· | ···················· | ····················· | ···················· | ···················· | ···················· |
| N | 5686 | hCoV-19/England/MILK-A476D4/2020\|EPI_ISL_598494\|20 | ···················· | ···················· | ····················· | ···················· | ···················· | ···················· |
| N | 5687 | hCoV-19/England/MILK-A470A9/2020\|EPI_ISL_598495\|20 | ···················· | ···················· | ····················· | ···················· | ···················· | ···················· |
| N | 5688 | hCoV-19/England/MILK-A47FD9/2020\|EPI_ISL_598496\|20 | ···················· | ···················· | ····················· | ···················· | ···················· | ···················· |
| N | 5689 | hCoV-19/England/MILK-A47E72/2020\|EPI_ISL_598497\|20 | ···················· | ·A·················· | ····················· | ···················· | ···················· | ···················· |
| N | 5690 | hCoV-19/England/MILK-A46D94/2020\|EPI_ISL_598498\|20 | ···················· | ···················· | ····················· | ···················· | ···················· | ···················· |
| N | 5691 | hCoV-19/England/MILK-A47C3C/2020\|EPI_ISL_598499\|20 | ···················· | ···················· | ····················· | ···················· | ···················· | ···················· |
| N | 5692 | hCoV-19/England/MILK-A4574C/2020\|EPI_ISL_598546\|20 | ···················· | ···················· | ····················· | ···················· | ···················· | ···················· |
| N | 5693 | hCoV-19/England/MILK-A44D0F/2020\|EPI_ISL_598547\|20 | ···················· | ···················· | ····················· | ···················· | ···················· | ···················· |
| N | 5694 | hCoV-19/England/MILK-A44EEE/2020\|EPI_ISL_598548\|20 | ···················· | ·A·················· | ····················· | ···················· | ···················· | ···················· |
| N | 5695 | hCoV-19/England/MILK-A459CE/2020\|EPI_ISL_598549\|20 | ···················· | ···················· | ····················· | ···················· | ···················· | ···················· |
| N | 5696 | hCoV-19/England/MILK-A45E83/2020\|EPI_ISL_598550\|20 | ···················· | ···················· | ····················· | ···················· | ···················· | ···················· |
| N | 5697 | hCoV-19/England/MILK-A44C20/2020\|EPI_ISL_598551\|20 | ···················· | ···················· | ····················· | ···················· | ···················· | ···················· |
| N | 5698 | hCoV-19/England/MILK-A44FFA/2020\|EPI_ISL_598552\|20 | ···················· | ···················· | ····················· | ···················· | ···················· | ···················· |
| N | 5699 | hCoV-19/England/MILK-A458FE/2020\|EPI_ISL_598553\|20 | ···················· | ···················· | ····················· | ···················· | ···················· | ···················· |
| N | 5700 | hCoV-19/England/MILK-A45A16/2020\|EPI_ISL_598554\|20 | ···················· | ···················· | ····················· | ···················· | ···················· | ···················· |
| N | 5701 | hCoV-19/England/MILK-A45AAD/2020\|EPI_ISL_598555\|20 | ···················· | ···················· | ····················· | ···················· | ···················· | ···················· |
| N | 5702 | hCoV-19/England/MILK-A45E0B/2020\|EPI_ISL_598562\|20 | ···················· | ···················· | ····················· | ···················· | ···················· | ···················· |
| N | 5703 | hCoV-19/England/MILK-A45B40/2020\|EPI_ISL_598563\|20 | ···················· | ···················· | ····················· | ···················· | ···················· | ···················· |
| N | 5704 | hCoV-19/England/MILK-A44D69/2020\|EPI_ISL_598566\|20 | ···················· | ···················· | ····················· | ···················· | ···················· | ···················· |
| N | 5705 | hCoV-19/England/MILK-A0689F/2020\|EPI_ISL_599876\|20 | ···················· | ···················· | ····················· | ···················· | ···················· | ···················· |

N 5706 hCoV-19/England/MILK-A05502/2020|EPI_ISL_599877|20 .................... .................... .................... .................... .................... ....................
N 5707 hCoV-19/Wales/MILK-A05669/2020|EPI_ISL_599878|2020 .................... .................... .................... .................... .................... ....................
N 5708 hCoV-19/England/MILK-A05FE6/2020|EPI_ISL_599879|20 .................... .................... .................... .................... .................... ....................
N 5709 hCoV-19/England/MILK-A05A21/2020|EPI_ISL_599880|20 .................... .................... .................... .................... .................... ....................
N 5710 hCoV-19/England/MILK-A06817/2020|EPI_ISL_599881|20 .................... .................... .................... .................... .................... ....................
N 5711 hCoV-19/Wales/MILK-A06440/2020|EPI_ISL_599882|2020 .................... .................... .................... .................... .................... ....................
N 5712 hCoV-19/England/MILK-A068BD/2020|EPI_ISL_599883|20 .................... .................... .................... .................... .................... ....................
N 5713 hCoV-19/Wales/MILK-A057FD/2020|EPI_ISL_599884|2020 .................... .................... .................... .................... .................... ....................
N 5714 hCoV-19/England/MILK-A067B0/2020|EPI_ISL_599885|20 .................... .................... .................... .................... .................... ....................
N 5715 hCoV-19/England/MILK-A0603D/2020|EPI_ISL_599890|20 .................... .................... .................... .................... .................... ....................
N 5716 hCoV-19/England/MILK-A06A4E/2020|EPI_ISL_599891|20 .................... .................... .................... .................... .................... ....................
N 5717 hCoV-19/England/MILK-A05678/2020|EPI_ISL_599892|20 .................... .................... .................... .................... .................... ....................
N 5718 hCoV-19/England/MILK-A06A99/2020|EPI_ISL_599893|20 .................... .................... .................... .................... .................... ....................
N 5719 hCoV-19/England/MILK-A0572A/2020|EPI_ISL_599894|20 .................... .................... .................... .................... .................... ....................
N 5720 hCoV-19/Wales/MILK-A05DDD/2020|EPI_ISL_599895|2020 .................... .................... .................... .................... .................... ....................
N 5721 hCoV-19/England/MILK-A062CE/2020|EPI_ISL_599897|20 .................... .A.................. .................... .................... .................... ..A.................
N 5722 hCoV-19/USA/NM-DOH-2020309753/2020|EPI_ISL_602200| .................... .A.................. .................... .................... .................... ....................
N 5723 hCoV-19/USA/NM-DOH-2020309771/2020|EPI_ISL_602201| .................... .................... .................... .................... .................... ....................
N 5724 hCoV-19/USA/NM-DOH-2020309772/2020|EPI_ISL_602202| .................... .................... .................... .................... .................... ....................
N 5725 hCoV-19/USA/NM-DOH-2020310165/2020|EPI_ISL_602203| .................... .A.................. .................... .................... .................... ....................
N 5726 hCoV-19/USA/NM-DOH-2020310604/2020|EPI_ISL_602204| .................... .................... .................... .................... .................... ....................
N 5727 hCoV-19/USA/NM-DOH-2020310635/2020|EPI_ISL_602205| .................... .................... .................... .................... .................... ....................
N 5728 hCoV-19/USA/NM-DOH-2020310691/2020|EPI_ISL_602206| .................... .A.................. .................... .................... .................... ....................
N 5729 hCoV-19/USA/NM-DOH-2020310774/2020|EPI_ISL_602207| .................... .................... .................... .................... .................... ....................
N 5730 hCoV-19/USA/NM-LANL-2020016075/2020|EPI_ISL_602208 .................... .................... .................... .................... .................... ....................
N 5731 hCoV-19/USA/NM-LANL-2020016077/2020|EPI_ISL_602209 .................... .................... .................... .................... .................... ....................
N 5732 hCoV-19/USA/MD-MDH-0235/2020|EPI_ISL_602258|2020-0 .................... .................... .................... .................... .................... ....................
N 5733 hCoV-19/USA/MD-MDH-0236/2020|EPI_ISL_602259|2020-0 .................... .............A...... .................... .................... .................... ....................
N 5734 hCoV-19/USA/MD-MDH-0237/2020|EPI_ISL_602260|2020-0 .................... .A.................. .................... .................... .................... ....................
N 5735 hCoV-19/USA/MD-MDH-0238/2020|EPI_ISL_602261|2020-0 .................... .................... .................... .................... .................... ....................
N 5736 hCoV-19/USA/MD-MDH-0239/2020|EPI_ISL_602262|2020-0 .................... .................... .................... .................... .................... ....................
N 5737 hCoV-19/USA/MD-MDH-0240/2020|EPI_ISL_602263|2020-1 .................... .................... .................... .................... .................... ....................
N 5738 hCoV-19/USA/MD-MDH-0241/2020|EPI_ISL_602264|2020-1 .................... .................... .................... .................... .................... ....................
N 5739 hCoV-19/USA/MD-MDH-0242/2020|EPI_ISL_602265|2020-1 .................... .................... .................... .................... .................... ....................
N 5740 hCoV-19/USA/MD-MDH-0243/2020|EPI_ISL_602266|2020-1 .................... .................... .................... .................... .................... ....................
N 5741 hCoV-19/USA/MD-MDH-0244/2020|EPI_ISL_602267|2020-1 .................... .................... .................... .................... .................... ....................
N 5742 hCoV-19/USA/MD-MDH-0245/2020|EPI_ISL_602268|2020-1 .................... .................... .................... .................... .................... ....................
N 5743 hCoV-19/USA/MD-MDH-0246/2020|EPI_ISL_602269|2020-1 .................... .................... .................... .................... .................... ....................
N 5744 hCoV-19/USA/MD-MDH-0247/2020|EPI_ISL_602270|2020-1 .................... .................... .................... .................... .................... ....................
N 5745 hCoV-19/USA/MD-MDH-0248/2020|EPI_ISL_602271|2020-1 .................... ............C....... .................... .................... .................... ....................
N 5746 hCoV-19/USA/MD-MDH-0249/2020|EPI_ISL_602272|2020-1 .................... .................... .................... .................... .................... ....................
N 5747 hCoV-19/USA/MD-MDH-0250/2020|EPI_ISL_602273|2020-1 .................... .................... .................... .................... .................... ....................
N 5748 hCoV-19/USA/MD-MDH-0251/2020|EPI_ISL_602274|2020-1 .................... .................... .................... .................... .................... ....................
N 5749 hCoV-19/USA/MD-MDH-0252/2020|EPI_ISL_602275|2020-1 .................... .................... .................... .................... .................... ....................
N 5750 hCoV-19/USA/MD-MDH-0253/2020|EPI_ISL_602276|2020-1 .................... .................... .................... .................... .................... ....................
N 5751 hCoV-19/USA/MD-MDH-0254/2020|EPI_ISL_602277|2020-1 .................... .A.................. .................... .................... .................... ....................
N 5752 hCoV-19/USA/NM-LANL-2020016089/2020|EPI_ISL_602278 .................... .................... .................... .................... .................... ....................
N 5753 hCoV-19/USA/NM-LANL-2020016415/2020|EPI_ISL_602279 .................... .................... .................... .................... .................... ....................
N 5754 hCoV-19/USA/NM-LANL-2020016419/2020|EPI_ISL_602280 .................... .................... .................... .................... .................... ....................
N 5755 hCoV-19/USA/NM-LANL-2020016449/2020|EPI_ISL_602281 .................... .................... .................... .................... .................... ....................
N 5756 hCoV-19/Germany/NW-UBI-1/2020|EPI_ISL_602282|2020- .................... .................... .................... .................... .................... ....................
N 5757 hCoV-19/Germany/NW-UBI-2/2020|EPI_ISL_602283|2020- .................... .................... .................... .................... .................... ....................
N 5758 hCoV-19/Germany/NW-UBI-3/2020|EPI_ISL_602284|2020- .................... .................... .................... .................... .................... ....................
N 5759 hCoV-19/Germany/NW-UBI-4/2020|EPI_ISL_602285|2020- .................... .................... .................... .................... .................... ....................
N 5760 hCoV-19/Germany/NW-UBI-5/2020|EPI_ISL_602286|2020- .................... .................... .................... .................... .................... ....................
N 5761 hCoV-19/Germany/NW-UBI-6/2020|EPI_ISL_602287|2020- .................... .................... .................... .................... .................... ....................
N 5762 hCoV-19/Germany/NW-UBI-7/2020|EPI_ISL_602288|2020- .................... .................... .................... .................... .................... ....................
N 5763 hCoV-19/Germany/NW-UBI-8/2020|EPI_ISL_602289|2020- .................... .................... .................... .................... .................... ....................
N 5764 hCoV-19/Germany/NW-UBI-9/2020|EPI_ISL_602290|2020- .................... .................... .................... .................... .................... ....................
N 5765 hCoV-19/Germany/NW-UBI-10/2020|EPI_ISL_602291|2020 .................... .................... .................... .................... .................... ....................
N 5766 hCoV-19/Germany/NW-UBI-11/2020|EPI_ISL_602292|2020 .................... .................... .................... .................... .................... ....................
N 5767 hCoV-19/Germany/NW-UBI-12/2020|EPI_ISL_602293|2020 .................... .................... .................... .................... .................... ....................
N 5768 hCoV-19/Germany/NW-UBI-13/2020|EPI_ISL_602294|2020 .................... .................... .................... .................... .................... ....................
N 5769 hCoV-19/Germany/NW-UBI-14/2020|EPI_ISL_602295|2020 .................... .................... .................... .................... .................... ....................
N 5770 hCoV-19/Germany/NW-UBI-15/2020|EPI_ISL_602296|2020 .................... .................... .................... .................... .................... ....................
N 5771 hCoV-19/Germany/NW-UBI-16/2020|EPI_ISL_602297|2020 .................... .................... .................... .................... .................... ....................
N 5772 hCoV-19/Germany/NW-UBI-17/2020|EPI_ISL_602298|2020 .................... .................... .................... .................... .................... ....................
N 5773 hCoV-19/Germany/NW-UBI-18/2020|EPI_ISL_602299|2020 .................... .................... .................... .................... .................... ....................
N 5774 hCoV-19/Switzerland/ZH-ETHZ-310432/2020|EPI_ISL_60 .................... .................... .................... A................... .................... ....................
N 5775 hCoV-19/Switzerland/ZH-ETHZ-310433/2020|EPI_ISL_60 .................... .................... .................... .................... .................... ....................
N 5776 hCoV-19/Switzerland/ZH-ETHZ-310434/2020|EPI_ISL_60 .................... .................... .................... .................... .................... ....................
N 5777 hCoV-19/Switzerland/ZH-ETHZ-310435/2020|EPI_ISL_60 .................... .................... .................... .................... .................... ....................
N 5778 hCoV-19/Switzerland/BE-ETHZ-310437/2020|EPI_ISL_60 .................... .................... .................... .................... .................... ....................
N 5779 hCoV-19/Switzerland/ZH-ETHZ-310438/2020|EPI_ISL_60 .................... .................... .................... .................... .................... ....................
N 5780 hCoV-19/Switzerland/BE-ETHZ-310440/2020|EPI_ISL_60 .................... .................... .................... .................... .................... ....................
N 5781 hCoV-19/Switzerland/ZH-ETHZ-310441/2020|EPI_ISL_60 .................... .................... .................... .................... .................... ....................
N 5782 hCoV-19/Switzerland/ZH-ETHZ-310442/2020|EPI_ISL_60 .................... .................... .................... .................... .................... ....................
N 5783 hCoV-19/Switzerland/BE-ETHZ-310443/2020|EPI_ISL_60 .................... .................... .................... .................... .................... ....................
N 5784 hCoV-19/Switzerland/ZH-ETHZ-310444/2020|EPI_ISL_60 .................... .................... .................... .................... .................... ....................
N 5785 hCoV-19/Switzerland/BE-ETHZ-310445/2020|EPI_ISL_60 .................... .................... .................... .................... .................... ....................

N 5786 hCoV-19/Switzerland/VD-ETHZ-310446/2020|EPI_ISL_60 ···················· ···················· ···················· ···················· ······K············· ····················
N 5787 hCoV-19/Switzerland/BE-ETHZ-310447/2020|EPI_ISL_60 ···················· ···················· ···················· ···················· ···················· ····················
N 5788 hCoV-19/Switzerland/BE-ETHZ-310448/2020|EPI_ISL_60 ···················· ···················· ···················· ···················· ···················· ····················
N 5789 hCoV-19/Switzerland/BE-ETHZ-310450/2020|EPI_ISL_60 ···················· ···················· ···················· ···················· ···················· ····················
N 5790 hCoV-19/Switzerland/VD-ETHZ-310451/2020|EPI_ISL_60 ···················· ···················· ···················· ···················· ···················· ····················
N 5791 hCoV-19/Switzerland/BE-ETHZ-310452/2020|EPI_ISL_60 ···················· ···················· ···················· ···················· ······K············· ····················
N 5792 hCoV-19/Switzerland/ZH-ETHZ-310453/2020|EPI_ISL_60 ···················· ···················· ···················· ···················· ······K············· ····················
N 5793 hCoV-19/Switzerland/TI-ETHZ-310454/2020|EPI_ISL_60 ···················· ···················· ···················· ···················· ···················· ····················
N 5794 hCoV-19/Switzerland/BE-ETHZ-310455/2020|EPI_ISL_60 ···················· ···················· ···················· ···················· ······K············· ····················
N 5795 hCoV-19/Switzerland/AR-ETHZ-310457/2020|EPI_ISL_60 ···················· ···················· ···················· ···················· ···················· ····················
N 5796 hCoV-19/Switzerland/SG-ETHZ-310459/2020|EPI_ISL_60 ···················· ···················· ···················· ···················· ···················· ····················
N 5797 hCoV-19/Switzerland/GR-ETHZ-310462/2020|EPI_ISL_60 ···················· ···················· ···················· ···················· ···················· ····················
N 5798 hCoV-19/Switzerland/BE-ETHZ-310464/2020|EPI_ISL_60 ···················· ···················· ···················· ···················· ···················· ····················
N 5799 hCoV-19/Switzerland/BE-ETHZ-310465/2020|EPI_ISL_60 ···················· ···················· ···················· ···················· ···················· ····················
N 5800 hCoV-19/Switzerland/BE-ETHZ-310467/2020|EPI_ISL_60 ···················· ···················· ···················· ···················· ······K············· ····················
N 5801 hCoV-19/Switzerland/UR-ETHZ-310469/2020|EPI_ISL_60 ···················· ···················· ···················· ···················· ···················· ····················
N 5802 hCoV-19/Switzerland/GE-ETHZ-310470/2020|EPI_ISL_60 ···················· ···················· ···················· ···················· ······K············· ····················
N 5803 hCoV-19/Switzerland/GE-ETHZ-310471/2020|EPI_ISL_60 ···················· ···················· ···················· ···················· ···················· ····················
N 5804 hCoV-19/Switzerland/GE-ETHZ-310472/2020|EPI_ISL_60 ···················· ···················· ···················· ···················· ···················· ····················
N 5805 hCoV-19/Switzerland/ZH-ETHZ-310473/2020|EPI_ISL_60 ···················· ···················· ···················· ···················· ···················· ····················
N 5806 hCoV-19/Switzerland/GE-ETHZ-310474/2020|EPI_ISL_60 ···················· ···················· ···················· ···················· ···················· ····················
N 5807 hCoV-19/Switzerland/BL-ETHZ-310477/2020|EPI_ISL_60 ···················· ···················· ···················· ···················· ···················· ····················
N 5808 hCoV-19/Switzerland/AG-ETHZ-310478/2020|EPI_ISL_60 ···················· ···················· ···················· ···················· ···················· ····················
N 5809 hCoV-19/Switzerland/BL-ETHZ-310479/2020|EPI_ISL_60 ···················· ···················· ···················· ···················· ···················· ····················
N 5810 hCoV-19/Switzerland/BE-ETHZ-310491/2020|EPI_ISL_60 ···················· ···················· ···················· ···················· ···················· ····················
N 5811 hCoV-19/Switzerland/VS-ETHZ-310492/2020|EPI_ISL_60 ···················· ···················· ···················· ···················· ···················· ····················
N 5812 hCoV-19/Switzerland/VS-ETHZ-310493/2020|EPI_ISL_60 ···················· ···················· ···················· ···················· ···················· ····················
N 5813 hCoV-19/Switzerland/VD-ETHZ-310502/2020|EPI_ISL_60 ···················· ···················· ···················· ···················· ···················· ····················
N 5814 hCoV-19/Switzerland/BE-ETHZ-310503/2020|EPI_ISL_60 ···················· ···················· ···················· ···················· ···················· ····················
N 5815 hCoV-19/Switzerland/BE-ETHZ-310504/2020|EPI_ISL_60 ···················· ···················· ···················· ···················· ···················· ····················
N 5816 hCoV-19/Switzerland/VD-ETHZ-310505/2020|EPI_ISL_60 ···················· ···················· ···················· ···················· ···················· ····················
N 5817 hCoV-19/Switzerland/VD-ETHZ-310506/2020|EPI_ISL_60 ···················· ···················· ···················· ···················· ···················· ····················
N 5818 hCoV-19/Switzerland/VD-ETHZ-310507/2020|EPI_ISL_60 ···················· ···················· ···················· ···················· ···················· ····················
N 5819 hCoV-19/Switzerland/VD-ETHZ-310508/2020|EPI_ISL_60 ···················· ···················· ···················· ···················· ···················· ····················
N 5820 hCoV-19/Switzerland/VS-ETHZ-310509/2020|EPI_ISL_60 ···················· ···················· ···················· ···················· ···················· ····················
N 5821 hCoV-19/Switzerland/BE-ETHZ-310510/2020|EPI_ISL_60 ···················· ···················· ···················· ···················· ···················· ····················
N 5822 hCoV-19/Switzerland/BE-ETHZ-310511/2020|EPI_ISL_60 ···················· ···················· ···················· ···················· ···················· ····················
N 5823 hCoV-19/Switzerland/BE-ETHZ-310512/2020|EPI_ISL_60 ···················· ···················· ···················· ···················· ···················· ····················
N 5824 hCoV-19/Switzerland/BE-ETHZ-310513/2020|EPI_ISL_60 ···················· ···················· ···················· ···················· ···················· ····················
N 5825 hCoV-19/Switzerland/GR-ETHZ-310514/2020|EPI_ISL_60 ···················· ···················· ···················· ···················· ······K············· ····················
N 5826 hCoV-19/Switzerland/BE-ETHZ-310515/2020|EPI_ISL_60 ···················· ···················· ···················· ···················· ···················· ····················
N 5827 hCoV-19/Switzerland/TI-ETHZ-310516/2020|EPI_ISL_60 ···················· ···················· ···················· ···················· ···················· ····················
N 5828 hCoV-19/Switzerland/TI-ETHZ-310517/2020|EPI_ISL_60 ···················· ···················· ···················· ···················· ···················· ····················
N 5829 hCoV-19/Switzerland/BE-ETHZ-310518/2020|EPI_ISL_60 ···················· ···················· ···················· ···················· ···················· ····················
N 5830 hCoV-19/Switzerland/BE-ETHZ-310519/2020|EPI_ISL_60 ···················· ···················· ···················· ···················· ···················· ····················
N 5831 hCoV-19/Switzerland/BE-ETHZ-310520/2020|EPI_ISL_60 ···················· ···················· ···················· ···················· ···················· ····················
N 5832 hCoV-19/Switzerland/GR-ETHZ-310521/2020|EPI_ISL_60 ···················· ···················· ···················· ···················· ······K············· ····················
N 5833 hCoV-19/Switzerland/BE-ETHZ-310522/2020|EPI_ISL_60 ···················· ···················· ···················· ···················· ···················· ····················
N 5834 hCoV-19/Switzerland/BE-ETHZ-310523/2020|EPI_ISL_60 ···················· ···················· ···················· ···················· ···················· ····················
N 5835 hCoV-19/Switzerland/BE-ETHZ-310524/2020|EPI_ISL_60 ···················· ···················· ···················· ···················· ···················· ····················
N 5836 hCoV-19/Switzerland/BE-ETHZ-310525/2020|EPI_ISL_60 ···················· ···················· ···················· ···················· ···················· ····················
N 5837 hCoV-19/Switzerland/BE-ETHZ-310526/2020|EPI_ISL_60 ···················· ···················· ···················· ···················· ···················· ····················
N 5838 hCoV-19/Switzerland/GE-ETHZ-310527/2020|EPI_ISL_60 ···················· ···················· ···················· ···················· ···················· ····················
N 5839 hCoV-19/Switzerland/BE-ETHZ-310528/2020|EPI_ISL_60 ···················· ···················· ···················· ···················· ···················· ····················
N 5840 hCoV-19/Switzerland/GE-ETHZ-310529/2020|EPI_ISL_60 ···················· ···················· ···················· ···················· ···················· ····················
N 5841 hCoV-19/Switzerland/BE-ETHZ-310530/2020|EPI_ISL_60 ···················· ···················· ···················· ···················· ···················· ····················
N 5842 hCoV-19/Switzerland/BE-ETHZ-310531/2020|EPI_ISL_60 ···················· ···················· ···················· ···················· ···················· ····················
N 5843 hCoV-19/USA/PA-QDX-2437/2020|EPI_ISL_604837|2020-0 ···················· ···················· ···················· ···················· ···················· ····················
N 5844 hCoV-19/USA/PA-QDX-2455/2020|EPI_ISL_604838|2020-0 ···················· ···················· ···················· ···················· ···················· ····················
N 5845 hCoV-19/USA/PA-QDX-2459/2020|EPI_ISL_604839|2020-0 ···················· ···················· ···················· ···················· ···················· ····················
N 5846 hCoV-19/USA/PA-QDX-2497/2020|EPI_ISL_604840|2020-0 ···················· ···················· ···················· ···················· ···················· ····················
N 5847 hCoV-19/USA/SC-QDX-2484/2020|EPI_ISL_604841|2020-0 ···················· ···················· ···················· ···················· ···················· ····················
N 5848 hCoV-19/USA/TN-QDX-2517/2020|EPI_ISL_604842|2020-0 ···················· ···················· ···················· ···················· ···················· ····················
N 5849 hCoV-19/USA/TX-QDX-2554/2020|EPI_ISL_604843|2020-0 ···················· ···················· ···················· ···················· ···················· ····················
N 5850 hCoV-19/USA/UT-QDX-2434/2020|EPI_ISL_604844|2020-0 ···················· ···················· ···················· ···················· ···················· ····················
N 5851 hCoV-19/USA/UT-QDX-2440/2020|EPI_ISL_604845|2020-0 ···················· ···················· ···················· ···················· ···················· ····················
N 5852 hCoV-19/USA/UT-QDX-2454/2020|EPI_ISL_604846|2020-0 ···················· ···················· ···················· ···················· ···················· ····················
N 5853 hCoV-19/USA/UT-QDX-2469/2020|EPI_ISL_604847|2020-0 ···················· ···················· ···················· ···················· ···················· ····················
N 5854 hCoV-19/USA/UT-QDX-2476/2020|EPI_ISL_604848|2020-0 ···················· ···················· ···················· ···················· ···················· ····················
N 5855 hCoV-19/USA/UT-QDX-2479/2020|EPI_ISL_604849|2020-0 ···················· ···················· ···················· ···················· ···················· ····················
N 5856 hCoV-19/USA/UT-QDX-2480/2020|EPI_ISL_604850|2020-0 ···················· ···················· ···················· ···················· ···················· ····················
N 5857 hCoV-19/USA/UT-QDX-2487/2020|EPI_ISL_604851|2020-0 ···················· ···················· ···················· ···················· ···················· ····················
N 5858 hCoV-19/USA/UT-QDX-2490/2020|EPI_ISL_604852|2020-0 ···················· ···················· ···················· ···················· ···················· ····················
N 5859 hCoV-19/USA/UT-QDX-2491/2020|EPI_ISL_604853|2020-0 ···················· ···················· ···················· ···················· ···················· ····················
N 5860 hCoV-19/USA/UT-QDX-2492/2020|EPI_ISL_604854|2020-0 ···················· ···················· ···················· ···················· ···················· ····················
N 5861 hCoV-19/USA/UT-QDX-2504/2020|EPI_ISL_604855|2020-0 ···················· ···················· ···················· ···················· ···················· ····················
N 5862 hCoV-19/USA/UT-QDX-2505/2020|EPI_ISL_604856|2020-0 ···················· ···················· ···················· ···················· ···················· ····················
N 5863 hCoV-19/USA/UT-QDX-2506/2020|EPI_ISL_604857|2020-0 ···················· ···················· ···················· ···················· ···················· ····················
N 5864 hCoV-19/USA/UT-QDX-2509/2020|EPI_ISL_604858|2020-0 ···················· ···················· ···················· ···················· ···················· ····················
N 5865 hCoV-19/USA/UT-QDX-2523/2020|EPI_ISL_604859|2020-0 ···················· ···················· ···················· ···················· ···················· ····················

| | | | | | | | | |
|---|---|---|---|---|---|---|---|---|
| N | 5866 | hCoV-19/USA/UT-QDX-2526/2020\|EPI_ISL_604860\|2020-0 | ···················· | ···················· | ···················· | ···················· | ···················· | ···················· |
| N | 5867 | hCoV-19/USA/UT-QDX-2527/2020\|EPI_ISL_604861\|2020-0 | ···················· | ···················· | ···················· | ···················· | ···················· | ···················· |
| N | 5868 | hCoV-19/USA/UT-QDX-2531/2020\|EPI_ISL_604862\|2020-0 | ···················· | ···················· | ···················· | ···················· | ···················· | ···················· |
| N | 5869 | hCoV-19/USA/UT-QDX-2532/2020\|EPI_ISL_604863\|2020-0 | ···················· | ···················· | ···················· | ···················· | ···················· | ···················· |
| N | 5870 | hCoV-19/USA/UT-QDX-2549/2020\|EPI_ISL_604864\|2020-0 | ···················· | ···················· | ···················· | ···················· | ···················· | ···················· |
| N | 5871 | hCoV-19/USA/UT-QDX-2550/2020\|EPI_ISL_604865\|2020-0 | ···················· | ···················· | ···················· | ···················· | ···················· | ···················· |
| N | 5872 | hCoV-19/USA/UT-QDX-2556/2020\|EPI_ISL_604866\|2020-0 | ···················· | ···················· | ···················· | ···················· | ···················· | ···················· |
| N | 5873 | hCoV-19/USA/UT-QDX-2557/2020\|EPI_ISL_604867\|2020-0 | ···················· | ···················· | ···················· | ···················· | ···················· | ···················· |
| N | 5874 | hCoV-19/USA/WA-QDX-2441/2020\|EPI_ISL_604868\|2020-0 | ···················· | ···················· | ···················· | ···················· | ···················· | ···················· |
| N | 5875 | hCoV-19/USA/WA-QDX-2471/2020\|EPI_ISL_604869\|2020-0 | ···················· | ···················· | ···················· | ···················· | ···················· | ···················· |
| N | 5876 | hCoV-19/USA/WA-QDX-2514/2020\|EPI_ISL_604870\|2020-0 | ···················· | ···················· | ···················· | ···················· | ···················· | ···················· |
| N | 5877 | hCoV-19/USA/WA-QDX-2542/2020\|EPI_ISL_604871\|2020-0 | ···················· | ···················· | ···················· | ···················· | ···················· | ···················· |
| N | 5878 | hCoV-19/USA/WA-QDX-2548/2020\|EPI_ISL_604872\|2020-0 | ···················· | ···················· | ···················· | ···················· | ···················· | ···················· |
| N | 5879 | hCoV-19/USA/AZ-QDX-2673/2020\|EPI_ISL_604873\|2020-0 | ···················· | ···················· | ···················· | ···················· | ···················· | ···················· |
| N | 5880 | hCoV-19/USA/CA-QDX-2570/2020\|EPI_ISL_604874\|2020-0 | ···················· | ···················· | ···················· | ···················· | ···················· | ···················· |
| N | 5881 | hCoV-19/USA/CA-QDX-2576/2020\|EPI_ISL_604875\|2020-0 | ···················· | ···················· | ···················· | ···················· | ···················· | ···················· |
| N | 5882 | hCoV-19/USA/CA-QDX-2578/2020\|EPI_ISL_604876\|2020-0 | ···················· | ···················· | ···················· | ···················· | ···················· | ···················· |
| N | 5883 | hCoV-19/USA/CA-QDX-2581/2020\|EPI_ISL_604877\|2020-0 | ···················· | ···················· | ···················· | ···················· | ···················· | ···················· |
| N | 5884 | hCoV-19/USA/CA-QDX-2583/2020\|EPI_ISL_604878\|2020-0 | ···················· | ···················· | ···················· | ···················· | ···················· | ···················· |
| N | 5885 | hCoV-19/USA/CA-QDX-2584/2020\|EPI_ISL_604879\|2020-0 | ···················· | ···················· | ···················· | ···················· | ···················· | ···················· |
| N | 5886 | hCoV-19/USA/CA-QDX-2603/2020\|EPI_ISL_604890\|2020-0 | ···················· | ···················· | ···················· | ···················· | ···················· | ···················· |
| N | 5887 | hCoV-19/USA/CA-QDX-2609/2020\|EPI_ISL_604893\|2020-0 | ···················· | ···················· | ···················· | ···················· | ···················· | ···················· |
| N | 5888 | hCoV-19/USA/CA-QDX-2610/2020\|EPI_ISL_604894\|2020-0 | ···················· | ···················· | ···················· | ···················· | ···················· | ···················· |
| N | 5889 | hCoV-19/USA/MI-UM-10036306710/2020\|EPI_ISL_596604\| | ···················· | ·············A······ | ···················· | ···················· | ···················· | ···················· |
| N | 5890 | hCoV-19/USA/MI-UM-10036309295/2020\|EPI_ISL_596605\| | ···················· | ···················· | ···················· | ···················· | ···················· | ···················· |
| N | 5891 | hCoV-19/England/CAMC-A57D09/2020\|EPI_ISL_597200\|20 | ···················· | ···················· | ···················· | ···················· | ···················· | ···················· |
| N | 5892 | hCoV-19/England/CAMC-A582CD/2020\|EPI_ISL_597201\|20 | ···················· | ···················· | ···················· | ···················· | ···················· | ···················· |
| N | 5893 | hCoV-19/England/CAMC-A57E60/2020\|EPI_ISL_597202\|20 | ···················· | ···················· | ···················· | ···················· | ···················· | ···················· |
| N | 5894 | hCoV-19/England/CAMC-A5910B/2020\|EPI_ISL_597203\|20 | ···················· | ···················· | ···················· | ···················· | ···················· | ···················· |
| N | 5895 | hCoV-19/England/CAMC-A58BD1/2020\|EPI_ISL_597204\|20 | ···················· | ···················· | ···················· | ···················· | ···················· | ···················· |
| N | 5896 | hCoV-19/England/CAMC-A57E42/2020\|EPI_ISL_597206\|20 | ···················· | ···················· | ···················· | ···················· | ···················· | ···················· |
| N | 5897 | hCoV-19/England/CAMC-A5961B/2020\|EPI_ISL_597207\|20 | ···················· | ···················· | ···················· | ···················· | ···················· | ···················· |
| N | 5898 | hCoV-19/England/CAMC-A585B5/2020\|EPI_ISL_597208\|20 | ···················· | ···················· | ···················· | ···················· | ···················· | ···················· |
| N | 5899 | hCoV-19/England/CAMC-A5849A/2020\|EPI_ISL_597209\|20 | ···················· | ···················· | ···················· | ···················· | ···················· | ···················· |
| N | 5900 | hCoV-19/England/CAMC-A586FE/2020\|EPI_ISL_597210\|20 | ···················· | ···················· | ···················· | ···················· | ···················· | ···················· |
| N | 5901 | hCoV-19/England/CAMC-A581C0/2020\|EPI_ISL_597211\|20 | ···················· | ···················· | ···················· | ···················· | ···················· | ···················· |
| N | 5902 | hCoV-19/England/CAMC-A58904/2020\|EPI_ISL_597212\|20 | ···················· | ···················· | ···················· | ···················· | ···················· | ···················· |
| N | 5903 | hCoV-19/England/CAMC-A57DFA/2020\|EPI_ISL_597213\|20 | ···················· | ···················· | ···················· | ···················· | ···················· | ···················· |
| N | 5904 | hCoV-19/England/CAMC-A58500/2020\|EPI_ISL_597225\|20 | ···················· | ···················· | ···················· | ···················· | ···················· | ···················· |
| N | 5905 | hCoV-19/England/CAMC-A592CC/2020\|EPI_ISL_597226\|20 | ···················· | ···················· | ···················· | ···················· | ···················· | ···················· |
| N | 5906 | hCoV-19/England/CAMC-A58F8A/2020\|EPI_ISL_597227\|20 | ···················· | ···················· | ···················· | ···················· | ···················· | ···················· |
| N | 5907 | hCoV-19/England/CAMC-A58922/2020\|EPI_ISL_597228\|20 | ···················· | ·A·················· | ···················· | ···················· | ···················· | ··A················· |
| N | 5908 | hCoV-19/England/CAMC-A58A98/2020\|EPI_ISL_597229\|20 | ···················· | ·A·················· | ···················· | ···················· | ···················· | ···················· |
| N | 5909 | hCoV-19/England/QEUH-A5E91D/2020\|EPI_ISL_597235\|20 | ···················· | ···················· | ···················· | ···················· | ···················· | ···················· |
| N | 5910 | hCoV-19/England/QEUH-A5E2C7/2020\|EPI_ISL_597236\|20 | ···················· | ···················· | ···················· | ···················· | ···················· | ···················· |
| N | 5911 | hCoV-19/England/CAMC-A57598/2020\|EPI_ISL_597237\|20 | ···················· | ···················· | ···················· | ···················· | ···················· | ···················· |
| N | 5912 | hCoV-19/England/CAMC-A57282/2020\|EPI_ISL_597238\|20 | ···················· | ···················· | ···················· | ···················· | ···················· | ···················· |
| N | 5913 | hCoV-19/England/CAMC-A57729/2020\|EPI_ISL_597239\|20 | ···················· | ···················· | ···················· | ···················· | ···················· | ···················· |
| N | 5914 | hCoV-19/England/QEUH-A5E124/2020\|EPI_ISL_597240\|20 | ···················· | ···················· | ···················· | ···················· | ···················· | ···················· |
| N | 5915 | hCoV-19/England/QEUH-A5E528/2020\|EPI_ISL_597242\|20 | ···················· | ···················· | ···················· | ···················· | ···················· | ···················· |
| N | 5916 | hCoV-19/England/CAMC-A574E6/2020\|EPI_ISL_597243\|20 | ···················· | ···················· | ···················· | ···················· | ······T············· | ···················· |
| N | 5917 | hCoV-19/England/QEUH-A5DF2B/2020\|EPI_ISL_597244\|20 | ···················· | ···················· | ···················· | ···················· | ···················· | ···················· |
| N | 5918 | hCoV-19/England/CAMC-A57088/2020\|EPI_ISL_597246\|20 | ···················· | ···················· | ···················· | ···················· | ···················· | ···················· |
| N | 5919 | hCoV-19/England/CAMC-A59C55/2020\|EPI_ISL_597247\|20 | ···················· | ···················· | ···················· | ···················· | ···················· | ···················· |
| N | 5920 | hCoV-19/Wales/CAMC-A570D3/2020\|EPI_ISL_597248\|2020 | ···················· | ···················· | ···················· | ···················· | ···················· | ···················· |
| N | 5921 | hCoV-19/England/QEUH-A5E045/2020\|EPI_ISL_597249\|20 | ···················· | ···················· | ···················· | ···················· | ···················· | ···················· |
| N | 5922 | hCoV-19/England/CAMC-A57B87/2020\|EPI_ISL_597250\|20 | ···················· | ···················· | ···················· | ···················· | ···················· | ···················· |
| N | 5923 | hCoV-19/England/QEUH-A5E670/2020\|EPI_ISL_597252\|20 | ···················· | ···················· | ···················· | ···················· | ···················· | ···················· |
| N | 5924 | hCoV-19/England/CAMC-A5703D/2020\|EPI_ISL_597253\|20 | ···················· | ···················· | ···················· | ···················· | ···················· | ···················· |
| N | 5925 | hCoV-19/England/CAMC-A57B78/2020\|EPI_ISL_597254\|20 | ···················· | ···················· | ···················· | ···················· | ···················· | ···················· |
| N | 5926 | hCoV-19/England/CAMC-A57B96/2020\|EPI_ISL_597255\|20 | ···················· | ···················· | ···················· | ···················· | ···················· | ···················· |
| N | 5927 | hCoV-19/England/CAMC-A575C5/2020\|EPI_ISL_597256\|20 | ···················· | ···················· | ···················· | ···················· | ···················· | ···················· |
| N | 5928 | hCoV-19/England/CAMC-A57219/2020\|EPI_ISL_597257\|20 | ···················· | ···················· | ···················· | ···················· | ······T············· | ···················· |
| N | 5929 | hCoV-19/England/CAMC-A579F6/2020\|EPI_ISL_597258\|20 | ···················· | ···················· | ···················· | ···················· | ···················· | ···················· |
| N | 5930 | hCoV-19/England/CAMC-A57A02/2020\|EPI_ISL_597259\|20 | ···················· | ···················· | ···················· | ···················· | ···················· | ··A················· |
| N | 5931 | hCoV-19/England/QEUH-A5E69E/2020\|EPI_ISL_597260\|20 | ···················· | ···················· | ···················· | ···················· | ···················· | ···················· |
| N | 5932 | hCoV-19/England/QEUH-A5E7B9/2020\|EPI_ISL_597261\|20 | ···················· | ···················· | ···················· | ···················· | ···················· | ···················· |
| N | 5933 | hCoV-19/England/CAMC-A571FE/2020\|EPI_ISL_597262\|20 | ···················· | ···················· | ···················· | ···················· | ···················· | ···················· |
| N | 5934 | hCoV-19/England/QEUH-A5E0FA/2020\|EPI_ISL_597263\|20 | ···················· | ···················· | ···················· | ···················· | ···················· | ···················· |
| N | 5935 | hCoV-19/England/CAMC-A5771A/2020\|EPI_ISL_597264\|20 | ···················· | ···················· | ···················· | ···················· | ···················· | ···················· |
| N | 5936 | hCoV-19/England/CAMC-A57079/2020\|EPI_ISL_597266\|20 | ···················· | ···················· | ···················· | ···················· | ···················· | ···················· |
| N | 5937 | hCoV-19/England/QEUH-A5E7D7/2020\|EPI_ISL_597267\|20 | ···················· | ···················· | ···················· | ···················· | ···················· | ···················· |
| N | 5938 | hCoV-19/England/QEUH-A5DF67/2020\|EPI_ISL_597268\|20 | ···················· | ···················· | ···················· | ···················· | ···················· | ···················· |
| N | 5939 | hCoV-19/England/CAMC-A56FB9/2020\|EPI_ISL_597270\|20 | ···················· | ···················· | ···················· | ···················· | ···················· | ···················· |
| N | 5940 | hCoV-19/England/QEUH-A5E652/2020\|EPI_ISL_597272\|20 | ···················· | ···················· | ···················· | ···················· | ···················· | ···················· |
| N | 5941 | hCoV-19/England/CAMC-A59C0A/2020\|EPI_ISL_597273\|20 | ···················· | ···················· | ···················· | ···················· | ···················· | ···················· |
| N | 5942 | hCoV-19/England/CAMC-A572EC/2020\|EPI_ISL_597274\|20 | ···················· | ···················· | ···················· | ···················· | ···················· | ···················· |
| N | 5943 | hCoV-19/England/QEUH-A5E4EF/2020\|EPI_ISL_597275\|20 | ···················· | ···················· | ···················· | ···················· | ···················· | ···················· |
| N | 5944 | hCoV-19/England/QEUH-A5DFD0/2020\|EPI_ISL_597276\|20 | ···················· | ···················· | ···················· | ···················· | ···················· | ···················· |
| N | 5945 | hCoV-19/England/QEUH-A5E31F/2020\|EPI_ISL_597278\|20 | ···················· | ···················· | ···················· | ···················· | ···················· | ···················· |

| | | | | | | | | |
|---|---|---|---|---|---|---|---|---|
| N | 5946 | hCoV-19/England/CAMC-A579C9/2020\|EPI_ISL_597279\|20 | ···················· | ···················· | ···················· | ···················· | ···················· | ···················· |
| N | 5947 | hCoV-19/England/ALDP-A5281C/2020\|EPI_ISL_597900\|20 | ···················· | ···················· | ···················· | ···················· | ···················· | ···················· |
| N | 5948 | hCoV-19/England/ALDP-A51FAF/2020\|EPI_ISL_597901\|20 | ···················· | ···················· | ···················· | ···················· | ···················· | ···················· |
| N | 5949 | hCoV-19/England/ALDP-A51385/2020\|EPI_ISL_597902\|20 | ···················· | ···················· | ···················· | ···················· | ···················· | ···················· |
| N | 5950 | hCoV-19/England/ALDP-A52472/2020\|EPI_ISL_597903\|20 | ···················· | ···················· | ···················· | ···················· | ···················· | ···················· |
| N | 5951 | hCoV-19/England/ALDP-A513EF/2020\|EPI_ISL_597904\|20 | ···················· | ···················· | ···················· | ···················· | ···················· | ···················· |
| N | 5952 | hCoV-19/England/ALDP-A516B9/2020\|EPI_ISL_597905\|20 | ···················· | ···················· | ···················· | ···················· | ···················· | ···················· |
| N | 5953 | hCoV-19/England/ALDP-A514DD/2020\|EPI_ISL_597906\|20 | ···················· | ·A·················· | ···················· | ···················· | ···················· | ··A················· |
| N | 5954 | hCoV-19/England/ALDP-A51E39/2020\|EPI_ISL_597907\|20 | ···················· | ···················· | ···················· | ···················· | ···················· | ···················· |
| N | 5955 | hCoV-19/England/ALDP-A528C1/2020\|EPI_ISL_597908\|20 | ···················· | ···················· | ···················· | ···················· | ···················· | ···················· |
| N | 5956 | hCoV-19/England/ALDP-A51561/2020\|EPI_ISL_597909\|20 | ···················· | ···················· | ···················· | ···················· | ···················· | ···················· |
| N | 5957 | hCoV-19/England/ALDP-A5274C/2020\|EPI_ISL_597938\|20 | ···················· | ···················· | ···················· | ···················· | ···················· | ···················· |
| N | 5958 | hCoV-19/England/MILK-A46CA6/2020\|EPI_ISL_598500\|20 | ···················· | ···················· | ···················· | ···················· | ···················· | ···················· |
| N | 5959 | hCoV-19/England/MILK-A4723A/2020\|EPI_ISL_598501\|20 | ···················· | ···················· | ···················· | ···················· | ···················· | ···················· |
| N | 5960 | hCoV-19/England/MILK-A47443/2020\|EPI_ISL_598502\|20 | ···················· | ···················· | ···················· | ···················· | ···················· | ···················· |
| N | 5961 | hCoV-19/England/MILK-A47CB4/2020\|EPI_ISL_598503\|20 | ···················· | ···················· | ···················· | ···················· | ···················· | ···················· |
| N | 5962 | hCoV-19/England/MILK-A47188/2020\|EPI_ISL_598504\|20 | ···················· | ···················· | ···················· | ···················· | ···················· | ···················· |
| N | 5963 | hCoV-19/England/MILK-A47294/2020\|EPI_ISL_598505\|20 | ···················· | ···················· | ···················· | ···················· | ···················· | ···················· |
| N | 5964 | hCoV-19/England/MILK-A46CD3/2020\|EPI_ISL_598507\|20 | ···················· | ···················· | ···················· | ···················· | ···················· | ···················· |
| N | 5965 | hCoV-19/England/MILK-A47DEE/2020\|EPI_ISL_598508\|20 | ···················· | ···················· | ···················· | ···················· | ···················· | ···················· |
| N | 5966 | hCoV-19/England/MILK-A47892/2020\|EPI_ISL_598509\|20 | ···················· | ···················· | ···················· | ···················· | ···················· | ···················· |
| N | 5967 | hCoV-19/England/MILK-A46EEC/2020\|EPI_ISL_598510\|20 | ···················· | ···················· | ···················· | ···················· | ···················· | ···················· |
| N | 5968 | hCoV-19/England/MILK-A47935/2020\|EPI_ISL_598511\|20 | ···················· | ···················· | ···················· | ···················· | ···················· | ···················· |
| N | 5969 | hCoV-19/England/MILK-A47522/2020\|EPI_ISL_598512\|20 | ···················· | ···················· | ···················· | ···················· | ···················· | ···················· |
| N | 5970 | hCoV-19/England/MILK-A47FE8/2020\|EPI_ISL_598513\|20 | ···················· | ···················· | ···················· | ···················· | ···················· | ···················· |
| N | 5971 | hCoV-19/England/MILK-A46E82/2020\|EPI_ISL_598515\|20 | ···················· | ···················· | ···················· | ···················· | ···················· | ···················· |
| N | 5972 | hCoV-19/England/MILK-A48002/2020\|EPI_ISL_598516\|20 | ···················· | ···················· | ···················· | ···················· | ···················· | ···················· |
| N | 5973 | hCoV-19/England/MILK-A47C96/2020\|EPI_ISL_598517\|20 | ···················· | ···················· | ···················· | ···················· | ···················· | ···················· |
| N | 5974 | hCoV-19/England/MILK-A47CE1/2020\|EPI_ISL_598518\|20 | ···················· | ···················· | ···················· | ···················· | ···················· | ···················· |
| N | 5975 | hCoV-19/England/MILK-A471F1/2020\|EPI_ISL_598519\|20 | ···················· | ···················· | ···················· | ···················· | ···················· | ···················· |
| N | 5976 | hCoV-19/England/MILK-A479CC/2020\|EPI_ISL_598520\|20 | ···················· | ···················· | ···················· | ···················· | ···················· | ···················· |
| N | 5977 | hCoV-19/England/MILK-A46ECE/2020\|EPI_ISL_598521\|20 | ···················· | ···················· | ···················· | ···················· | ···················· | ···················· |
| N | 5978 | hCoV-19/England/MILK-A478FC/2020\|EPI_ISL_598522\|20 | ···················· | ···················· | ···················· | ···················· | ···················· | ···················· |
| N | 5979 | hCoV-19/England/MILK-A4807B/2020\|EPI_ISL_598523\|20 | ···················· | ···················· | ···················· | ···················· | ···················· | ···················· |
| N | 5980 | hCoV-19/England/MILK-A461A7/2020\|EPI_ISL_598525\|20 | ···················· | ···················· | ···················· | ···················· | ···················· | ···················· |
| N | 5981 | hCoV-19/England/MILK-A4621D/2020\|EPI_ISL_598526\|20 | ···················· | ·A·················· | ···················· | ···················· | ···················· | ··A················· |
| N | 5982 | hCoV-19/England/MILK-A457E2/2020\|EPI_ISL_598527\|20 | ···················· | ···················· | ···················· | ···················· | ···················· | ···················· |
| N | 5983 | hCoV-19/England/MILK-A45C6B/2020\|EPI_ISL_598528\|20 | ···················· | ···················· | ···················· | ···················· | ···················· | ···················· |
| N | 5984 | hCoV-19/England/MILK-A464AE/2020\|EPI_ISL_598529\|20 | ···················· | ···················· | ···················· | ···················· | ···················· | ···················· |
| N | 5985 | hCoV-19/England/MILK-A457F1/2020\|EPI_ISL_598531\|20 | ···················· | ···················· | ···················· | ···················· | ···················· | ···················· |
| N | 5986 | hCoV-19/England/MILK-A45788/2020\|EPI_ISL_598533\|20 | ···················· | ·A·················· | ···················· | ···················· | ···················· | ··A················· |
| N | 5987 | hCoV-19/England/MILK-A45E47/2020\|EPI_ISL_598534\|20 | ···················· | ···················· | ···················· | ···················· | ···················· | ···················· |
| N | 5988 | hCoV-19/England/MILK-A46277/2020\|EPI_ISL_598536\|20 | ···················· | ·A·················· | ···················· | ···················· | ···················· | ···················· |
| N | 5989 | hCoV-19/England/MILK-A45BF5/2020\|EPI_ISL_598537\|20 | ···················· | ···················· | ···················· | ···················· | ···················· | ···················· |
| N | 5990 | hCoV-19/England/MILK-A466F3/2020\|EPI_ISL_598539\|20 | ···················· | ···················· | ···················· | ···················· | A··················· | ···················· |
| N | 5991 | hCoV-19/England/MILK-A44B50/2020\|EPI_ISL_598540\|20 | ···················· | ···················· | ···················· | ···················· | ···················· | ···················· |
| N | 5992 | hCoV-19/England/MILK-A45C7A/2020\|EPI_ISL_598541\|20 | ···················· | ···················· | ···················· | ···················· | ···················· | ···················· |
| N | 5993 | hCoV-19/England/MILK-A4575B/2020\|EPI_ISL_598542\|20 | ···················· | ···················· | ···················· | ···················· | ···················· | ···················· |
| N | 5994 | hCoV-19/England/MILK-A45CE3/2020\|EPI_ISL_598543\|20 | ···················· | ···················· | ···················· | ···················· | ···················· | ···················· |
| N | 5995 | hCoV-19/England/MILK-A462FF/2020\|EPI_ISL_598544\|20 | ···················· | ···················· | ···················· | ···················· | ···················· | ···················· |
| N | 5996 | hCoV-19/England/MILK-A45B7D/2020\|EPI_ISL_598556\|20 | ···················· | ···················· | ···················· | ···················· | ···················· | ···················· |
| N | 5997 | hCoV-19/England/MILK-A44C7B/2020\|EPI_ISL_598557\|20 | ···················· | ···················· | ···················· | ···················· | ···················· | ···················· |
| N | 5998 | hCoV-19/England/MILK-A458C1/2020\|EPI_ISL_598558\|20 | ···················· | ···················· | ···················· | ···················· | ···················· | ···················· |
| N | 5999 | hCoV-19/England/MILK-A44D3C/2020\|EPI_ISL_598559\|20 | ···················· | ···················· | ···················· | ···················· | ···················· | ···················· |
| N | 6000 | hCoV-19/England/MILK-A44B7E/2020\|EPI_ISL_598567\|20 | ···················· | ···················· | ···················· | ···················· | ·····T·············· | ···················· |
| N | 6001 | hCoV-19/England/MILK-A45885/2020\|EPI_ISL_598569\|20 | ···················· | ·A·················· | ···················· | ···················· | ···················· | ··A················· |
| N | 6002 | hCoV-19/England/MILK-A44A62/2020\|EPI_ISL_598570\|20 | ···················· | ···················· | ···················· | ···················· | ···················· | ···················· |
| N | 6003 | hCoV-19/England/MILK-A44F90/2020\|EPI_ISL_598571\|20 | ···················· | ···················· | ···················· | ···················· | ···················· | ···················· |
| N | 6004 | hCoV-19/England/MILK-A468C0/2020\|EPI_ISL_598573\|20 | ···················· | ···················· | ···················· | ···················· | ···················· | ···················· |
| N | 6005 | hCoV-19/England/MILK-A44CB7/2020\|EPI_ISL_598575\|20 | ···················· | ···················· | ···················· | ···················· | ···················· | ···················· |
| N | 6006 | hCoV-19/England/MILK-A45B9B/2020\|EPI_ISL_598576\|20 | ···················· | ···················· | ···················· | ···················· | ···················· | ···················· |
| N | 6007 | hCoV-19/England/MILK-A4613E/2020\|EPI_ISL_598577\|20 | ···················· | ···················· | ···················· | ···················· | ···················· | ···················· |
| N | 6008 | hCoV-19/England/MILK-A45014/2020\|EPI_ISL_598578\|20 | ···················· | ···················· | ···················· | ···················· | ···················· | ···················· |
| N | 6009 | hCoV-19/England/MILK-A4606E/2020\|EPI_ISL_598579\|20 | ···················· | ·A·················· | ···················· | ···················· | ···················· | ··A················· |
| N | 6010 | hCoV-19/England/MILK-A45D86/2020\|EPI_ISL_598580\|20 | ···················· | ···················· | ···················· | ···················· | ···················· | ···················· |
| N | 6011 | hCoV-19/England/MILK-A46286/2020\|EPI_ISL_598581\|20 | ···················· | ·A·················· | ···················· | ···················· | ···················· | ···················· |
| N | 6012 | hCoV-19/England/MILK-A45E65/2020\|EPI_ISL_598582\|20 | ···················· | ···················· | ···················· | ···················· | ···················· | ···················· |
| N | 6013 | hCoV-19/England/MILK-A45F62/2020\|EPI_ISL_598583\|20 | ···················· | ···················· | ···················· | ···················· | ·····T·············· | ···················· |
| N | 6014 | hCoV-19/England/MILK-A45BAA/2020\|EPI_ISL_598584\|20 | ···················· | ·A·················· | ···················· | ···················· | ···················· | ··A················· |
| N | 6015 | hCoV-19/England/MILK-A466A8/2020\|EPI_ISL_598585\|20 | ···················· | ···················· | ···················· | ···················· | ···················· | ···················· |
| N | 6016 | hCoV-19/England/MILK-A46866/2020\|EPI_ISL_598586\|20 | ···················· | ···················· | ···················· | ···················· | ·····T·············· | ···················· |
| N | 6017 | hCoV-19/England/MILK-A46602/2020\|EPI_ISL_598587\|20 | ···················· | ···················· | ···················· | ···················· | ···················· | ···················· |
| N | 6018 | hCoV-19/England/MILK-A44FEB/2020\|EPI_ISL_598590\|20 | ···················· | ···················· | ···················· | ···················· | ···················· | ···················· |
| N | 6019 | hCoV-19/England/MILK-A45A8F/2020\|EPI_ISL_598591\|20 | ···················· | ···················· | ···················· | ···················· | ···················· | ···················· |
| N | 6020 | hCoV-19/England/MILK-A44DB4/2020\|EPI_ISL_598592\|20 | ···················· | ···················· | ···················· | ···················· | ···················· | ···················· |
| N | 6021 | hCoV-19/England/MILK-A461D4/2020\|EPI_ISL_598593\|20 | ···················· | ···················· | ···················· | ···················· | ···················· | ···················· |
| N | 6022 | hCoV-19/England/MILK-A44BC9/2020\|EPI_ISL_598594\|20 | ···················· | ·A·················· | ···················· | ···················· | ···················· | ··A················· |
| N | 6023 | hCoV-19/England/MILK-A45858/2020\|EPI_ISL_598595\|20 | ···················· | ···················· | ···················· | ···················· | ···················· | ···················· |
| N | 6024 | hCoV-19/England/MILK-A45032/2020\|EPI_ISL_598596\|20 | ···················· | ···················· | ···················· | ···················· | ···················· | ···················· |
| N | 6025 | hCoV-19/England/MILK-A46769/2020\|EPI_ISL_598598\|20 | ···················· | ···················· | ···················· | ···················· | ···················· | ···················· |

| | | | | | | | |
|---|---|---|---|---|---|---|---|
| N | 6026 hCoV-19/England/MILK-A467A5/2020\|EPI_ISL_598599\|20 | ···················· | ···················· | ····················· | ···················· | ···················· | ···················· |
| N | 6027 hCoV-19/England/QEUH-9DE10F/2020\|EPI_ISL_599800\|20 | ········T··········· | ···················· | ····················· | ···················· | ···················· | ···················· |
| N | 6028 hCoV-19/England/QEUH-9DE0E4/2020\|EPI_ISL_599801\|20 | ···················· | ···················· | ····················· | ···················· | ···················· | ···················· |
| N | 6029 hCoV-19/England/QEUH-9DDC5A/2020\|EPI_ISL_599802\|20 | ···················· | ···················· | ····················· | ···················· | ···················· | ···················· |
| N | 6030 hCoV-19/England/QEUH-9DDDA2/2020\|EPI_ISL_599804\|20 | ···················· | ···················· | ····················· | ···················· | ···················· | ···················· |
| N | 6031 hCoV-19/England/QEUH-9DE406/2020\|EPI_ISL_599805\|20 | ···················· | ···················· | ····················· | ···················· | ···················· | ···················· |
| N | 6032 hCoV-19/England/QEUH-9DD62F/2020\|EPI_ISL_599806\|20 | ···················· | ···················· | ····················· | ···················· | ···················· | ···················· |
| N | 6033 hCoV-19/Scotland/QEUH-9DD7FF/2020\|EPI_ISL_599807\|2 | ···················· | ···················· | ····················· | ···················· | ···················· | ···················· |
| N | 6034 hCoV-19/England/QEUH-9DD5F5/2020\|EPI_ISL_599808\|20 | ···················· | ···················· | ····················· | ···················· | ···················· | ···················· |
| N | 6035 hCoV-19/England/QEUH-9DCD1C/2020\|EPI_ISL_599809\|20 | ···················· | ···················· | ····················· | ···················· | ···················· | ···················· |
| N | 6036 hCoV-19/England/QEUH-9DDA6F/2020\|EPI_ISL_599810\|20 | ···················· | ···················· | ····················· | ···················· | ···················· | ···················· |
| N | 6037 hCoV-19/England/QEUH-9DD1E2/2020\|EPI_ISL_599813\|20 | ···················· | ···················· | ····················· | ···················· | ···················· | ···················· |
| N | 6038 hCoV-19/England/QEUH-9DCD0D/2020\|EPI_ISL_599815\|20 | ···················· | ···················· | ····················· | ···················· | ···················· | ···················· |
| N | 6039 hCoV-19/England/QEUH-9DD373/2020\|EPI_ISL_599817\|20 | ···················· | ···················· | ····················· | ···················· | ···················· | ···················· |
| N | 6040 hCoV-19/England/MILK-A05A12/2020\|EPI_ISL_599818\|20 | ···················· | ···················· | ····················· | ···················· | ···················· | ···················· |
| N | 6041 hCoV-19/England/MILK-A067FC/2020\|EPI_ISL_599819\|20 | ···················· | ···················· | ····················· | ···················· | ···················· | ···················· |
| N | 6042 hCoV-19/England/MILK-A05EE9/2020\|EPI_ISL_599820\|20 | ···················· | ···················· | ····················· | ···················· | ···················· | ···················· |
| N | 6043 hCoV-19/England/MILK-A05890/2020\|EPI_ISL_599822\|20 | ···················· | ···················· | ····················· | ···················· | ···················· | ···················· |
| N | 6044 hCoV-19/England/MILK-A05CFE/2020\|EPI_ISL_599823\|20 | ···················· | ···················· | ····················· | ···················· | ···················· | ···················· |
| N | 6045 hCoV-19/England/MILK-A06659/2020\|EPI_ISL_599824\|20 | ···················· | ···················· | ····················· | ···················· | ······T············· | ···················· |
| N | 6046 hCoV-19/England/MILK-A061FE/2020\|EPI_ISL_599825\|20 | ···················· | ·A·················· | ····················· | ···················· | ···················· | ···A················ |
| N | 6047 hCoV-19/England/MILK-A05450/2020\|EPI_ISL_599826\|20 | ···················· | ···················· | ····················· | ···················· | ···················· | ···················· |
| N | 6048 hCoV-19/England/MILK-A058BE/2020\|EPI_ISL_599827\|20 | ···················· | ···················· | ····················· | ···················· | ···················· | ···················· |
| N | 6049 hCoV-19/England/MILK-A05A6D/2020\|EPI_ISL_599828\|20 | ···················· | ···················· | ····················· | ···················· | ···················· | ···················· |
| N | 6050 hCoV-19/England/MILK-A05C2B/2020\|EPI_ISL_599829\|20 | ···················· | ···················· | ····················· | ···················· | ···················· | ···················· |
| N | 6051 hCoV-19/England/MILK-A062EC/2020\|EPI_ISL_599830\|20 | ···················· | ···················· | ····················· | ···················· | ······T············· | ···················· |
| N | 6052 hCoV-19/Wales/MILK-A06A02/2020\|EPI_ISL_599831\|2020 | ···················· | ···················· | ····················· | ···················· | ···················· | ···················· |
| N | 6053 hCoV-19/England/MILK-A05F13/2020\|EPI_ISL_599833\|20 | ···················· | ···················· | ····················· | ···················· | ···················· | ···················· |
| N | 6054 hCoV-19/Wales/MILK-A06598/2020\|EPI_ISL_599835\|2020 | ···················· | ···················· | ····················· | ···················· | ···················· | ···················· |
| N | 6055 hCoV-19/England/MILK-A067CF/2020\|EPI_ISL_599836\|20 | ···················· | ···················· | ····················· | ···················· | ···················· | ···················· |
| N | 6056 hCoV-19/England/MILK-A05B5B/2020\|EPI_ISL_599837\|20 | ···················· | ·A·················· | ····················· | ···················· | ···················· | ···A················ |
| N | 6057 hCoV-19/England/MILK-A056F0/2020\|EPI_ISL_599838\|20 | ···················· | ···················· | ····················· | ···················· | ···················· | ···················· |
| N | 6058 hCoV-19/Wales/MILK-A06589/2020\|EPI_ISL_599839\|2020 | ···················· | ···················· | ····················· | ···················· | ···················· | ···················· |
| N | 6059 hCoV-19/England/MILK-A055B7/2020\|EPI_ISL_599840\|20 | ···················· | ···················· | ····················· | ···················· | ···················· | ···················· |
| N | 6060 hCoV-19/England/MILK-A059BB/2020\|EPI_ISL_599841\|20 | ···················· | ···················· | ····················· | ···················· | ···················· | ···················· |
| N | 6061 hCoV-19/England/MILK-A060B5/2020\|EPI_ISL_599842\|20 | ···················· | ···················· | ····················· | ···················· | ···················· | ···················· |
| N | 6062 hCoV-19/England/MILK-A062FB/2020\|EPI_ISL_599843\|20 | ···················· | ···················· | ····················· | ···················· | ······T············· | ···················· |
| N | 6063 hCoV-19/England/MILK-A06765/2020\|EPI_ISL_599844\|20 | ···················· | ···················· | ····················· | ···················· | ···················· | ···················· |
| N | 6064 hCoV-19/England/MILK-A06871/2020\|EPI_ISL_599846\|20 | ···················· | ···················· | ····················· | ···················· | ···················· | ···················· |
| N | 6065 hCoV-19/Wales/MILK-A05827/2020\|EPI_ISL_599847\|2020 | ···················· | ···················· | ····················· | ···················· | ···················· | ···················· |
| N | 6066 hCoV-19/England/MILK-A059D9/2020\|EPI_ISL_599848\|20 | ···················· | ···················· | ····················· | ···················· | ···················· | ···················· |
| N | 6067 hCoV-19/England/MILK-A053BD/2020\|EPI_ISL_599849\|20 | ···················· | ···················· | ····················· | ···················· | ···················· | ···················· |
| N | 6068 hCoV-19/England/MILK-A063AD/2020\|EPI_ISL_599850\|20 | ···················· | ·A·················· | ····················· | ···················· | ···················· | ···A················ |
| N | 6069 hCoV-19/England/MILK-A06352/2020\|EPI_ISL_599851\|20 | ···················· | ···················· | ····················· | ···················· | ···················· | ···················· |
| N | 6070 hCoV-19/England/MILK-A06756/2020\|EPI_ISL_599852\|20 | ···················· | ···················· | ····················· | ···················· | ···················· | ···················· |
| N | 6071 hCoV-19/England/MILK-A06774/2020\|EPI_ISL_599853\|20 | ···················· | ···················· | ····················· | ···················· | ···················· | ···················· |
| N | 6072 hCoV-19/England/MILK-A067A1/2020\|EPI_ISL_599854\|20 | ···················· | ·A·················· | ····················· | ···················· | ···················· | ···················· |
| N | 6073 hCoV-19/England/MILK-A05C49/2020\|EPI_ISL_599855\|20 | ···················· | ···················· | ····················· | ···················· | ···················· | ···················· |
| N | 6074 hCoV-19/England/MILK-A06158/2020\|EPI_ISL_599856\|20 | ···················· | ···················· | ····················· | ···················· | ···················· | ···················· |
| N | 6075 hCoV-19/England/MILK-A059F7/2020\|EPI_ISL_599857\|20 | ···················· | ···················· | ····················· | ···················· | ···················· | ···················· |
| N | 6076 hCoV-19/England/MILK-A05D37/2020\|EPI_ISL_599858\|20 | ···················· | ···················· | ····················· | ···················· | ···················· | ···················· |
| N | 6077 hCoV-19/England/MILK-A0553F/2020\|EPI_ISL_599859\|20 | ···················· | ···················· | ····················· | ···················· | ···················· | ···················· |
| N | 6078 hCoV-19/England/MILK-A05766/2020\|EPI_ISL_599860\|20 | ···················· | ···················· | ····················· | ···················· | ···················· | ···················· |
| N | 6079 hCoV-19/England/MILK-A06738/2020\|EPI_ISL_599861\|20 | ···················· | ···················· | ····················· | ···················· | ···················· | ···················· |
| N | 6080 hCoV-19/England/MILK-A05EAD/2020\|EPI_ISL_599862\|20 | ···················· | ···················· | ····················· | ···················· | ···················· | ···················· |
| N | 6081 hCoV-19/England/MILK-A0638F/2020\|EPI_ISL_599863\|20 | ···················· | ···················· | ····················· | ···················· | ···················· | ···················· |
| N | 6082 hCoV-19/England/MILK-A05ECB/2020\|EPI_ISL_599864\|20 | ···················· | ···················· | ····················· | ···················· | ···················· | ···················· |
| N | 6083 hCoV-19/England/MILK-A0557B/2020\|EPI_ISL_599865\|20 | ···················· | ···················· | ····················· | ···················· | ···················· | ···················· |
| N | 6084 hCoV-19/England/MILK-A05F40/2020\|EPI_ISL_599866\|20 | ···················· | ···················· | ····················· | ···················· | ···················· | ···················· |
| N | 6085 hCoV-19/England/MILK-A055A8/2020\|EPI_ISL_599868\|20 | ···················· | ···················· | ····················· | ···················· | ···················· | ···················· |
| N | 6086 hCoV-19/England/MILK-A060A6/2020\|EPI_ISL_599869\|20 | ···················· | ···················· | ····················· | ···················· | ···················· | ···················· |
| N | 6087 hCoV-19/England/MILK-A05C76/2020\|EPI_ISL_599870\|20 | ···················· | ···················· | ····················· | ···················· | ···················· | ···················· |
| N | 6088 hCoV-19/England/MILK-A05FD7/2020\|EPI_ISL_599871\|20 | ···················· | ···················· | ····················· | ···················· | ···················· | ···················· |
| N | 6089 hCoV-19/England/MILK-A05AF4/2020\|EPI_ISL_599872\|20 | ···················· | ···················· | ····················· | ···················· | ···················· | ···················· |
| N | 6090 hCoV-19/England/MILK-A05FC8/2020\|EPI_ISL_599873\|20 | ···················· | ·A·················· | ····················· | ···················· | ···················· | ···················· |
| N | 6091 hCoV-19/England/MILK-A0563C/2020\|EPI_ISL_599874\|20 | ···················· | ···················· | ····················· | ···················· | ···················· | ···················· |
| N | 6092 hCoV-19/England/MILK-A05E52/2020\|EPI_ISL_599875\|20 | ···················· | ···················· | ····················· | ···················· | ···················· | ···················· |
| N | 6093 hCoV-19/England/MILK-A059E8/2020\|EPI_ISL_599887\|20 | ···················· | ···················· | ····················· | ···················· | ···················· | ···················· |
| N | 6094 hCoV-19/England/MILK-A0671A/2020\|EPI_ISL_599889\|20 | ···················· | ···················· | ····················· | ···················· | ···················· | ···················· |
| N | 6095 hCoV-19/England/MILK-A05775/2020\|EPI_ISL_599896\|20 | ···················· | ·A·················· | ····················· | ···················· | ···················· | ···A················ |
| N | 6096 hCoV-19/England/MILK-A05B2E/2020\|EPI_ISL_599898\|20 | ···················· | ·A·················· | ····················· | ···················· | ···················· | ···················· |
| N | 6097 hCoV-19/England/MILK-A058DC/2020\|EPI_ISL_599899\|20 | ···················· | ·A·······A·········· | ····················· | ···················· | ···················· | ···················· |
| N | 6098 hCoV-19/Scotland/QEUH-9FF645/2020\|EPI_ISL_600365\|2 | ···················· | ···················· | ····················· | ···················· | ···················· | ···················· |
| N | 6099 hCoV-19/England/QEUH-9FF45A/2020\|EPI_ISL_600366\|20 | ···················· | ···················· | ····················· | ···················· | ···················· | ···················· |
| N | 6100 hCoV-19/Wales/MILK-9D4C35/2020\|EPI_ISL_601696\|2020 | ···················· | ···················· | ····················· | ···················· | ···················· | ···················· |
| N | 6101 hCoV-19/England/MILK-9D3F00/2020\|EPI_ISL_601697\|20 | ···················· | ·A·················· | ····················· | ···················· | ···················· | ···A················ |
| N | 6102 hCoV-19/Switzerland/ZH-ETHZ-310426/2020\|EPI_ISL_60 | ···················· | ···················· | ····················· | ···················· | ···················· | ···················· |
| N | 6103 hCoV-19/Switzerland/BE-ETHZ-310427/2020\|EPI_ISL_60 | ···················· | ···················· | ····················· | ···················· | ···················· | ···················· |
| N | 6104 hCoV-19/Switzerland/ZH-ETHZ-310428/2020\|EPI_ISL_60 | ···················· | ···················· | ····················· | ···················· | ···················· | ···················· |
| N | 6105 hCoV-19/Switzerland/ZH-ETHZ-310429/2020\|EPI_ISL_60 | ···················· | ···················· | ····················· | ···················· | ···················· | ···················· |

| | | | | | | | | |
|---|---|---|---|---|---|---|---|---|
| N | 6106 | hCoV-19/Switzerland/SG-ETHZ-310430/2020\|EPI_ISL_60 | ···················· | ···················· | ····················· | ···················· | ···················· | ···················· |
| N | 6107 | hCoV-19/Switzerland/ZH-ETHZ-310431/2020\|EPI_ISL_60 | ···················· | ···················· | ····················· | ···················· | ···················· | ···················· |
| N | 6108 | hCoV-19/USA/NY-QDX-2446/2020\|EPI_ISL_604800\|2020-0 | ···················· | ···················· | ····················· | ···················· | ···················· | ···················· |
| N | 6109 | hCoV-19/USA/NY-QDX-2447/2020\|EPI_ISL_604801\|2020-0 | ···················· | ···················· | ····················· | ···················· | ···················· | ···················· |
| N | 6110 | hCoV-19/USA/NY-QDX-2451/2020\|EPI_ISL_604802\|2020-0 | ···················· | ···················· | ····················· | ···················· | ···················· | ···················· |
| N | 6111 | hCoV-19/USA/NY-QDX-2452/2020\|EPI_ISL_604803\|2020-0 | ···················· | ···················· | ····················· | ···················· | ···················· | ···················· |
| N | 6112 | hCoV-19/USA/NY-QDX-2453/2020\|EPI_ISL_604804\|2020-0 | ···················· | ···················· | ····················· | ···················· | ···················· | ···················· |
| N | 6113 | hCoV-19/USA/NY-QDX-2463/2020\|EPI_ISL_604805\|2020-0 | ···················· | ···················· | ····················· | ···················· | ···················· | ···················· |
| N | 6114 | hCoV-19/USA/NY-QDX-2464/2020\|EPI_ISL_604806\|2020-0 | ···················· | ···················· | ····················· | ···················· | ···················· | ···················· |
| N | 6115 | hCoV-19/USA/NY-QDX-2465/2020\|EPI_ISL_604807\|2020-0 | ···················· | ···················· | ····················· | ···················· | ···················· | ···················· |
| N | 6116 | hCoV-19/USA/NY-QDX-2466/2020\|EPI_ISL_604808\|2020-0 | ···················· | ···················· | ····················· | ···················· | ···················· | ···················· |
| N | 6117 | hCoV-19/USA/NY-QDX-2467/2020\|EPI_ISL_604809\|2020-0 | ···················· | ···················· | ····················· | ···················· | ···················· | ···················· |
| N | 6118 | hCoV-19/USA/NY-QDX-2473/2020\|EPI_ISL_604810\|2020-0 | ···················· | ···················· | ····················· | ···················· | ···················· | ···················· |
| N | 6119 | hCoV-19/USA/NY-QDX-2474/2020\|EPI_ISL_604811\|2020-0 | ···················· | ···················· | ····················· | ···················· | ···················· | ···················· |
| N | 6120 | hCoV-19/USA/NY-QDX-2481/2020\|EPI_ISL_604812\|2020-0 | ···················· | ···················· | ····················· | ···················· | ···················· | ···················· |
| N | 6121 | hCoV-19/USA/NY-QDX-2482/2020\|EPI_ISL_604813\|2020-0 | ···················· | ···················· | ····················· | ···················· | ···················· | ···················· |
| N | 6122 | hCoV-19/USA/NY-QDX-2483/2020\|EPI_ISL_604814\|2020-0 | ···················· | ···················· | ····················· | ···················· | ···················· | ···················· |
| N | 6123 | hCoV-19/USA/NY-QDX-2486/2020\|EPI_ISL_604815\|2020-0 | ···················· | ···················· | ····················· | ···················· | ···················· | ···················· |
| N | 6124 | hCoV-19/USA/NY-QDX-2494/2020\|EPI_ISL_604816\|2020-0 | ···················· | ···················· | ····················· | ···················· | ···················· | ···················· |
| N | 6125 | hCoV-19/USA/NY-QDX-2498/2020\|EPI_ISL_604817\|2020-0 | ···················· | ···················· | ····················· | ···················· | ···················· | ···················· |
| N | 6126 | hCoV-19/USA/NY-QDX-2499/2020\|EPI_ISL_604818\|2020-0 | ···················· | ···················· | ····················· | ···················· | ···················· | ···················· |
| N | 6127 | hCoV-19/USA/NY-QDX-2501/2020\|EPI_ISL_604819\|2020-0 | ···················· | ···················· | ····················· | ···················· | ···················· | ···················· |
| N | 6128 | hCoV-19/USA/NY-QDX-2508/2020\|EPI_ISL_604820\|2020-0 | ···················· | ···················· | ····················· | ···················· | ···················· | ···················· |
| N | 6129 | hCoV-19/USA/NY-QDX-2512/2020\|EPI_ISL_604821\|2020-0 | ···················· | ···················· | ····················· | ···················· | ···················· | ···················· |
| N | 6130 | hCoV-19/USA/NY-QDX-2521/2020\|EPI_ISL_604822\|2020-0 | ···················· | ···················· | ····················· | ···················· | ···················· | ···················· |
| N | 6131 | hCoV-19/USA/NY-QDX-2522/2020\|EPI_ISL_604823\|2020-0 | ···················· | ···················· | ····················· | ···················· | ···················· | ···················· |
| N | 6132 | hCoV-19/USA/NY-QDX-2528/2020\|EPI_ISL_604824\|2020-0 | ···················· | ···················· | ····················· | ···················· | ···················· | ···················· |
| N | 6133 | hCoV-19/USA/NY-QDX-2529/2020\|EPI_ISL_604825\|2020-0 | ···················· | ···················· | ····················· | ···················· | ···················· | ···················· |
| N | 6134 | hCoV-19/USA/NY-QDX-2530/2020\|EPI_ISL_604826\|2020-0 | ···················· | ···················· | ····················· | ···················· | ···················· | ···················· |
| N | 6135 | hCoV-19/USA/NY-QDX-2533/2020\|EPI_ISL_604827\|2020-0 | ···················· | ···················· | ····················· | ···················· | ···················· | ···················· |
| N | 6136 | hCoV-19/USA/NY-QDX-2534/2020\|EPI_ISL_604828\|2020-0 | ···················· | ···················· | ····················· | ···················· | ···················· | ···················· |
| N | 6137 | hCoV-19/USA/NY-QDX-2535/2020\|EPI_ISL_604829\|2020-0 | ···················· | ···················· | ····················· | ···················· | ···················· | ···················· |
| N | 6138 | hCoV-19/USA/NY-QDX-2536/2020\|EPI_ISL_604830\|2020-0 | ···················· | ···················· | ····················· | ···················· | ···················· | ···················· |
| N | 6139 | hCoV-19/USA/NY-QDX-2539/2020\|EPI_ISL_604831\|2020-0 | ···················· | ···················· | ····················· | ···················· | ···················· | ···················· |
| N | 6140 | hCoV-19/USA/NY-QDX-2543/2020\|EPI_ISL_604832\|2020-0 | ···················· | ···················· | ····················· | ···················· | ···················· | ···················· |
| N | 6141 | hCoV-19/USA/NY-QDX-2545/2020\|EPI_ISL_604833\|2020-0 | ···················· | ···················· | ····················· | ···················· | ···················· | ···················· |
| N | 6142 | hCoV-19/USA/NY-QDX-2546/2020\|EPI_ISL_604834\|2020-0 | ···················· | ···················· | ····················· | ···················· | ···················· | ···················· |
| N | 6143 | hCoV-19/USA/PA-QDX-2435/2020\|EPI_ISL_604835\|2020-0 | ···················· | ···················· | ····················· | ···················· | ···················· | ···················· |
| N | 6144 | hCoV-19/USA/PA-QDX-2436/2020\|EPI_ISL_604836\|2020-0 | ···················· | ···················· | ····················· | ···················· | ···················· | ···················· |
| N | 6145 | hCoV-19/USA/MI-UM-10036335546/2020\|EPI_ISL_596601\| | ···················· | ···················· | ····················· | ···················· | ···················· | ···················· |
| N | 6146 | hCoV-19/USA/MI-UM-10036284225/2020\|EPI_ISL_596602\| | ···················· | ···················· | ····················· | ···················· | ···················· | ···················· |
| N | 6147 | hCoV-19/USA/MI-UM-10036310651/2020\|EPI_ISL_596603\| | ···················· | ·············A······ | ····················· | ···················· | ···················· | ···················· |
| N | 6149 | hCoV-19/USA/MI-UM-10036287712/2020\|EPI_ISL_596608\| | ···················· | ·············A······ | ····················· | ···················· | ···················· | ···················· |
| N | 6150 | hCoV-19/USA/MI-UM-10036333448/2020\|EPI_ISL_596609\| | ···················· | ···················· | ····················· | ···················· | ···················· | ···················· |
| N | 6154 | hCoV-19/USA/MI-UM-10036324614/2020\|EPI_ISL_596614\| | ···················· | ···················· | ····················· | ···················· | ···················· | ···················· |
| N | 6155 | hCoV-19/USA/MI-UM-10036335555/2020\|EPI_ISL_596615\| | ···················· | ···················· | ····················· | ···················· | ···················· | ···················· |
| N | 6156 | hCoV-19/USA/MI-UM-10036372963/2020\|EPI_ISL_596616\| | ···················· | ·············A······ | ····················· | ···················· | ···················· | ···················· |
| N | 6157 | hCoV-19/USA/MI-UM-10036339633/2020\|EPI_ISL_596617\| | ···················· | ···················· | ····················· | ···················· | ···················· | ···················· |
| N | 6158 | hCoV-19/USA/MI-UM-10036370482/2020\|EPI_ISL_596618\| | ···················· | ·············A······ | ····················· | ···················· | ···················· | ···················· |
| N | 6159 | hCoV-19/USA/MI-UM-10036361683/2020\|EPI_ISL_596619\| | ···················· | ···················· | ····················· | ···················· | ···················· | ···················· |
| N | 6160 | hCoV-19/Ireland/D-SVUH-R6408/2020\|EPI_ISL_596645\|2 | ···················· | ···················· | ····················· | ···················· | ···················· | ···················· |
| N | 6161 | hCoV-19/Ireland/D-SVUH-R5890/2020\|EPI_ISL_596646\|2 | ···················· | ···················· | ····················· | ···················· | ···················· | ···················· |
| N | 6162 | hCoV-19/Ireland/D-SVUH-R5391/2020\|EPI_ISL_596648\|2 | ···················· | ···················· | ····················· | ···················· | ···················· | ···················· |
| N | 6163 | hCoV-19/Ireland/D-SVUH-R6519/2020\|EPI_ISL_596650\|2 | ···················· | ···················· | ····················· | ···················· | ···················· | ···················· |
| N | 6164 | hCoV-19/Ireland/D-SVUH-R8382/2020\|EPI_ISL_596651\|2 | ···················· | ···················· | ····················· | ···················· | ···················· | ···················· |
| N | 6165 | hCoV-19/Ireland/D-SVUH-R7175/2020\|EPI_ISL_596652\|2 | ···················· | ···················· | ····················· | ···················· | ···················· | ···················· |
| N | 6166 | hCoV-19/Ireland/D-SVUH-R6370/2020\|EPI_ISL_596654\|2 | ···················· | ···················· | ····················· | ···················· | ···················· | ···················· |
| N | 6167 | hCoV-19/Ireland/D-SVUH-R6989/2020\|EPI_ISL_596655\|2 | ···················· | ···················· | ····················· | ···················· | ···················· | ···················· |
| N | 6168 | hCoV-19/Ireland/D-SVUH-R6565/2020\|EPI_ISL_596659\|2 | ···················· | ···················· | ····················· | ···················· | ···················· | ···················· |
| N | 6169 | hCoV-19/Ireland/D-SVUH-R6716/2020\|EPI_ISL_596660\|2 | ···················· | ···················· | ····················· | ···················· | ···················· | ···················· |
| N | 6170 | hCoV-19/Ireland/D-SVUH-R6717/2020\|EPI_ISL_596661\|2 | ···················· | ···················· | ····················· | ···················· | ···················· | ···················· |
| N | 6171 | hCoV-19/Ireland/D-SVUH-R7377/2020\|EPI_ISL_596663\|2 | ···················· | ···················· | ····················· | ···················· | ···················· | ···················· |
| N | 6172 | hCoV-19/England/ALDP-A51297/2020\|EPI_ISL_597910\|20 | ···················· | ···················· | ····················· | ···················· | ···················· | ···················· |
| N | 6173 | hCoV-19/England/ALDP-A51482/2020\|EPI_ISL_597911\|20 | ···················· | ·A·················· | ····················· | ···················· | ···················· | ···················· |
| N | 6174 | hCoV-19/England/ALDP-A5207E/2020\|EPI_ISL_597912\|20 | ···················· | ···················· | ····················· | ···················· | ···················· | ···················· |
| N | 6175 | hCoV-19/England/ALDP-A5175C/2020\|EPI_ISL_597913\|20 | ···················· | ···················· | ····················· | ···················· | ···················· | ···················· |
| N | 6176 | hCoV-19/England/ALDP-A51F27/2020\|EPI_ISL_597914\|20 | ···················· | ···················· | ····················· | ···················· | ···················· | ···················· |
| N | 6177 | hCoV-19/England/ALDP-A511B8/2020\|EPI_ISL_597915\|20 | ···················· | ···················· | ····················· | ···················· | ···················· | ···················· |
| N | 6178 | hCoV-19/England/ALDP-A5116D/2020\|EPI_ISL_597916\|20 | ···················· | ···················· | ····················· | ···················· | ···················· | ···················· |
| N | 6179 | hCoV-19/England/ALDP-A522E1/2020\|EPI_ISL_597917\|20 | ···················· | ···················· | ····················· | ···················· | ···················· | ···················· |
| N | 6180 | hCoV-19/England/ALDP-A528B2/2020\|EPI_ISL_597918\|20 | ···················· | ·A·················· | ····················· | ···················· | ···················· | ··A················· |
| N | 6181 | hCoV-19/England/ALDP-A522A5/2020\|EPI_ISL_597919\|20 | ···················· | ···················· | ····················· | ···················· | ···················· | ···················· |
| N | 6182 | hCoV-19/England/ALDP-A51E57/2020\|EPI_ISL_597920\|20 | ···················· | ···················· | ····················· | ···················· | ···················· | ···················· |
| N | 6183 | hCoV-19/England/ALDP-A515AD/2020\|EPI_ISL_597921\|20 | ···················· | ···················· | ····················· | ···················· | ···················· | ···················· |
| N | 6184 | hCoV-19/England/ALDP-A51701/2020\|EPI_ISL_597923\|20 | ···················· | ···················· | ····················· | ···················· | ···················· | ···················· |
| N | 6185 | hCoV-19/England/ALDP-A525F7/2020\|EPI_ISL_597924\|20 | ···················· | ·A·················· | ····················· | ···················· | ···················· | ··A················· |
| N | 6186 | hCoV-19/England/ALDP-A525E8/2020\|EPI_ISL_597925\|20 | ···················· | ·A·················· | ····················· | ···················· | ···················· | ··A················· |
| N | 6187 | hCoV-19/England/ALDP-A516F5/2020\|EPI_ISL_597926\|20 | ···················· | ···················· | ····················· | ···················· | ···················· | ···················· |
| N | 6188 | hCoV-19/England/ALDP-A52454/2020\|EPI_ISL_597927\|20 | ···················· | ···················· | ····················· | ···················· | ···················· | ···················· |
| N | 6189 | hCoV-19/England/ALDP-A5276A/2020\|EPI_ISL_597928\|20 | ···················· | ···················· | ····················· | ···················· | ···················· | ···················· |

```
N 6190 hCoV-19/England/ALDP-A522D2/2020|EPI_ISL_597930|20 ···················· ···················· ···················· ···················· ···················· ····················
N 6191 hCoV-19/England/ALDP-A51F90/2020|EPI_ISL_597931|20 ···················· ···················· ···················· ···················· ···················· ····················
N 6192 hCoV-19/England/ALDP-A5268B/2020|EPI_ISL_597932|20 ···················· ···················· ···················· ···················· ···················· ····················
N 6193 hCoV-19/England/ALDP-A51D5A/2020|EPI_ISL_597933|20 ···················· ···················· ···················· ···················· ···················· ····················
N 6194 hCoV-19/England/ALDP-A51464/2020|EPI_ISL_597934|20 ···················· ···················· ···················· ···················· ···················· ····················
N 6195 hCoV-19/England/ALDP-A51604/2020|EPI_ISL_597935|20 ···················· ···················· ···················· ···················· ···················· ····················
N 6196 hCoV-19/England/ALDP-A51288/2020|EPI_ISL_597936|20 ···················· ···················· ···················· ···················· ···················· ····················
N 6197 hCoV-19/England/ALDP-A5177A/2020|EPI_ISL_597939|20 ···················· ···················· ···················· ···················· ···················· ····················
N 6198 hCoV-19/England/ALDP-A51394/2020|EPI_ISL_597940|20 ···················· ···················· ···················· ···················· ···················· ····················
N 6199 hCoV-19/England/ALDP-A51112/2020|EPI_ISL_597941|20 ···················· ·A·······A·········· ···················· ···················· ···················· ····················
N 6200 hCoV-19/England/ALDP-A52788/2020|EPI_ISL_597942|20 ···················· ···················· ···················· ···················· ···················· ····················
N 6201 hCoV-19/England/ALDP-A5206F/2020|EPI_ISL_597943|20 ···················· ···················· ···················· ···················· ···················· ····················
N 6202 hCoV-19/England/ALDP-A52630/2020|EPI_ISL_597944|20 ···················· ···················· ···················· ···················· ···················· ····················
N 6203 hCoV-19/England/ALDP-A51D78/2020|EPI_ISL_597945|20 ···················· ···················· ···················· ···················· ···················· ····················
N 6204 hCoV-19/England/ALDP-A510F7/2020|EPI_ISL_597946|20 ···················· ···················· ···················· ···················· ······T············· ····················
N 6205 hCoV-19/England/ALDP-A525D9/2020|EPI_ISL_597947|20 ···················· ···················· ···················· ···················· ···················· ····················
N 6206 hCoV-19/England/ALDP-A5216C/2020|EPI_ISL_597948|20 ···················· ···················· ···················· ···················· ···················· ····················
N 6207 hCoV-19/England/ALDP-A5118B/2020|EPI_ISL_597950|20 ···················· ···················· ···················· ···················· ···················· ····················
N 6208 hCoV-19/England/ALDP-A51534/2020|EPI_ISL_597973|20 ···················· ···················· ···················· ···················· ···················· ····················
N 6209 hCoV-19/England/ALDP-A526F4/2020|EPI_ISL_597974|20 ···················· ···················· ···················· ···················· ···················· ····················
N 6210 hCoV-19/England/ALDP-A51A9F/2020|EPI_ISL_597975|20 ···················· ···················· ···················· ···················· ···················· ····················
N 6211 hCoV-19/England/ALDP-A524BE/2020|EPI_ISL_597976|20 ···················· ···················· ···················· ···················· ···················· ····················
N 6212 hCoV-19/England/ALDP-A52779/2020|EPI_ISL_597978|20 ···················· ···················· ···················· ···················· ···················· ····················
N 6213 hCoV-19/England/ALDP-A517E3/2020|EPI_ISL_597979|20 ···················· ···················· ···················· ···················· ···················· ····················
N 6214 hCoV-19/England/ALDP-A5223C/2020|EPI_ISL_597980|20 ···················· ···················· ···················· ···················· ···················· ····················
N 6215 hCoV-19/England/ALDP-A513FE/2020|EPI_ISL_597981|20 ···················· ···················· ···················· ···················· ···················· ····················
N 6216 hCoV-19/England/ALDP-A52005/2020|EPI_ISL_597982|20 ···················· ···················· ···················· ···················· ···················· ····················
N 6217 hCoV-19/England/ALDP-A51E84/2020|EPI_ISL_597983|20 ···················· ···················· ···················· ···················· ···················· ····················
N 6218 hCoV-19/England/ALDP-A511D6/2020|EPI_ISL_597990|20 ···················· ···················· ···················· ···················· ···················· ····················
N 6219 hCoV-19/England/ALDP-A510BB/2020|EPI_ISL_597991|20 ···················· ···················· ···················· ···················· ···················· ····················
N 6220 hCoV-19/England/ALDP-A517C5/2020|EPI_ISL_597992|20 ···················· ···················· ···················· ···················· ···················· ····················
N 6221 hCoV-19/England/ALDP-A51E66/2020|EPI_ISL_597993|20 ···················· ···················· ···················· ···················· ···················· ····················
N 6222 hCoV-19/England/ALDP-A51EA2/2020|EPI_ISL_597994|20 ···················· ···················· ···················· ···················· ···················· ····················
N 6223 hCoV-19/Scotland/QEUH-A00394/2020|EPI_ISL_600316|2 ···················· ···················· ···················· ···················· ···················· ····················
N 6224 hCoV-19/England/QEUH-9FF88B/2020|EPI_ISL_600317|20 ···················· ···················· ···················· ···················· ···················· ····················
N 6225 hCoV-19/Scotland/QEUH-9FFE7A/2020|EPI_ISL_600318|2 ···················· ···················· ···················· ···················· ···················· ····················
N 6226 hCoV-19/England/QEUH-9FFBDD/2020|EPI_ISL_600319|20 ···················· ···················· ···················· ···················· ···················· ····················
N 6227 hCoV-19/Scotland/QEUH-9FFA3A/2020|EPI_ISL_600320|2 ···················· ···················· ···················· ···················· ···················· ····················
N 6228 hCoV-19/England/QEUH-9FF241/2020|EPI_ISL_600321|20 ···················· ···················· ···················· ···················· ····T··············· ····················
N 6229 hCoV-19/England/QEUH-A00112/2020|EPI_ISL_600322|20 ···················· ···················· ···················· ···················· ···················· ····················
N 6230 hCoV-19/England/QEUH-9FF5B1/2020|EPI_ISL_600323|20 ···················· ···················· ···················· ···················· ···················· ····················
N 6231 hCoV-19/England/QEUH-A00130/2020|EPI_ISL_600324|20 ···················· ···················· ···················· ···················· ···················· ····················
N 6232 hCoV-19/England/QEUH-9FFDB9/2020|EPI_ISL_600325|20 ···················· ·A·················· ···················· ···················· ···················· ···A················
N 6233 hCoV-19/England/QEUH-9FFB55/2020|EPI_ISL_600326|20 ···················· ···················· ···················· ···················· ···················· ····················
N 6234 hCoV-19/England/QEUH-9FFFC2/2020|EPI_ISL_600327|20 ···················· ·A·················· ···················· ···················· ···················· ····················
N 6235 hCoV-19/England/QEUH-9FF84F/2020|EPI_ISL_600328|20 ···················· ·A·················· ···················· ···················· ···················· ····················
N 6236 hCoV-19/Scotland/QEUH-9FF7E8/2020|EPI_ISL_600329|2 ···················· ···················· ···················· ···················· ···················· ····················
N 6237 hCoV-19/England/QEUH-9FFB73/2020|EPI_ISL_600330|20 ···················· ···················· ···················· ···················· ···················· ····················
N 6238 hCoV-19/England/QEUH-9FF180/2020|EPI_ISL_600331|20 ···················· ···················· ···················· ···················· ···················· ····················
N 6239 hCoV-19/England/QEUH-A008B3/2020|EPI_ISL_600332|20 ···················· ···················· ···················· ···················· ···················· ····················
N 6240 hCoV-19/Scotland/QEUH-A003D0/2020|EPI_ISL_600333|2 ···················· ···················· ···················· ···················· ···················· ····················
N 6241 hCoV-19/Scotland/QEUH-9FF85E/2020|EPI_ISL_600335|2 ···················· ···················· ···················· ···················· ···················· ····················
N 6242 hCoV-19/England/QEUH-9FF5FD/2020|EPI_ISL_600336|20 ···················· ···················· ···················· ···················· ···················· ····················
N 6243 hCoV-19/England/QEUH-A00015/2020|EPI_ISL_600337|20 ···················· ···················· ···················· ···················· ···················· ····················
N 6244 hCoV-19/England/QEUH-9FF320/2020|EPI_ISL_600360|20 ···················· ·A·················· ···················· ···················· ···················· ····················
N 6245 hCoV-19/England/QEUH-A00868/2020|EPI_ISL_600361|20 ···················· ·A·················· ···················· ···················· ···················· ···A················
N 6246 hCoV-19/Scotland/QEUH-9FF86D/2020|EPI_ISL_600362|2 ···················· ···················· ···················· ···················· ···················· ····················
N 6247 hCoV-19/England/QEUH-A00006/2020|EPI_ISL_600363|20 ···················· ···················· ···················· ···················· ···················· ····················
N 6248 hCoV-19/England/QEUH-9FF1BD/2020|EPI_ISL_600364|20 ···················· ···················· ···················· ···················· ···················· ····················
N 6249 hCoV-19/England/QEUH-A00033/2020|EPI_ISL_600367|20 ···················· ···················· ···················· ···················· ···················· ····················
N 6250 hCoV-19/England/QEUH-9FF1F9/2020|EPI_ISL_600368|20 ···················· ···················· ···················· ···················· ···················· ····················
N 6251 hCoV-19/England/QEUH-A003EF/2020|EPI_ISL_600371|20 ···················· ···················· ···················· ···················· ···················· ····················
N 6252 hCoV-19/England/QEUH-9FFFB3/2020|EPI_ISL_600372|20 ···················· ···················· ···················· ···················· ···················· ····················
N 6253 hCoV-19/England/QEUH-9FF36C/2020|EPI_ISL_600373|20 ···················· ···················· ···················· ···················· ······T············· ····················
N 6254 hCoV-19/England/QEUH-9FF3F3/2020|EPI_ISL_600375|20 ···················· ···················· ···················· ···················· ···················· ····················
N 6255 hCoV-19/Scotland/QEUH-A007F2/2020|EPI_ISL_600376|2 ···················· ···················· ···················· ···················· ···················· ····················
N 6256 hCoV-19/England/QEUH-9FFDD7/2020|EPI_ISL_600378|20 ···················· ···················· ···················· ···················· ···················· ····················
N 6257 hCoV-19/England/QEUH-A000CA/2020|EPI_ISL_600379|20 ···················· ···················· ···················· ···················· ···················· ····················
N 6258 hCoV-19/England/QEUH-A006B9/2020|EPI_ISL_600380|20 ···················· ···················· ···················· ···················· ···················· ····················
N 6259 hCoV-19/England/QEUH-9FF9B5/2020|EPI_ISL_600381|20 ···················· ···················· ···················· ···················· ···················· ····················
N 6260 hCoV-19/England/QEUH-A0030D/2020|EPI_ISL_600382|20 ···················· ···················· ···················· ···················· ···················· ····················
N 6261 hCoV-19/Scotland/QEUH-A0075C/2020|EPI_ISL_600384|2 ···················· ···················· ···················· ···················· ···················· ····················
N 6262 hCoV-19/England/QEUH-9FFD8C/2020|EPI_ISL_600385|20 ···················· ···················· ···················· ···················· ···················· ····················
N 6263 hCoV-19/England/QEUH-9FFDAA/2020|EPI_ISL_600389|20 ···················· ···················· ···················· ···················· ···················· ····················
N 6264 hCoV-19/England/QEUH-A00464/2020|EPI_ISL_600390|20 ···················· ···················· ···················· ···················· ···················· ····················
N 6265 hCoV-19/England/QEUH-9FF4F0/2020|EPI_ISL_600391|20 ···················· ···················· ···················· ···················· ···················· ····················
N 6266 hCoV-19/England/QEUH-9FFD7D/2020|EPI_ISL_600392|20 ···················· ···················· ···················· ···················· ···················· ····················
N 6267 hCoV-19/Scotland/QEUH-9FF6AF/2020|EPI_ISL_600394|2 ···················· ···················· ···················· ···················· ···················· ····················
N 6268 hCoV-19/England/QEUH-A0024C/2020|EPI_ISL_600396|20 ···················· ···················· ···················· ···················· ···················· ····················
N 6269 hCoV-19/England/QEUH-9FF2C9/2020|EPI_ISL_600397|20 ···················· ···················· ···················· ···················· ···················· ····················
```

| | | | | | | | | | |
|---|---|---|---|---|---|---|---|---|---|
| N | 6270 | hCoV-19/England/QEUH-9FF19F/2020\|EPI_ISL_600398\|20 | ···················· | ···················· | ···················· | ···················· | ···················· | ···················· | |
| N | 6271 | hCoV-19/England/QEUH-9FFD13/2020\|EPI_ISL_600399\|20 | ···················· | ···················· | ···················· | ···················· | ···················· | ···················· | |
| N | 6272 | hCoV-19/England/MILK-9D452B/2020\|EPI_ISL_601601\|20 | ···················· | ···················· | ···················· | ···················· | ···················· | ···················· | |
| N | 6273 | hCoV-19/England/MILK-9D3D8E/2020\|EPI_ISL_601602\|20 | ···················· | ···················· | ···················· | ···················· | ···················· | ···················· | |
| N | 6274 | hCoV-19/England/MILK-9D4F87/2020\|EPI_ISL_601647\|20 | ···················· | ···················· | ···················· | ···················· | ···················· | ···················· | |
| N | 6275 | hCoV-19/England/CAMC-9D2244/2020\|EPI_ISL_601648\|20 | ···················· | ···················· | ···················· | ···················· | ···················· | ···················· | |
| N | 6276 | hCoV-19/England/MILK-9D3C72/2020\|EPI_ISL_601649\|20 | ···················· | ···················· | ···················· | ···················· | ···················· | ···················· | |
| N | 6277 | hCoV-19/England/MILK-9D4EF3/2020\|EPI_ISL_601651\|20 | ···················· | ···················· | ···················· | ···················· | ···················· | ···················· | |
| N | 6278 | hCoV-19/England/MILK-9D4B1A/2020\|EPI_ISL_601653\|20 | ···················· | ···················· | ···················· | ···················· | ···················· | ···················· | |
| N | 6279 | hCoV-19/England/MILK-9D4039/2020\|EPI_ISL_601654\|20 | ···················· | ···················· | ···················· | ···················· | ···················· | ···················· | |
| N | 6280 | hCoV-19/Wales/MILK-9D4BFC/2020\|EPI_ISL_601655\|2020 | ···················· | ···················· | ···················· | ···················· | ···················· | ···················· | |
| N | 6281 | hCoV-19/England/MILK-9D4B0B/2020\|EPI_ISL_601656\|20 | ···················· | ····A··············· | ···················· | ···················· | ···················· | ···················· | |
| N | 6282 | hCoV-19/England/CAMC-9D22EA/2020\|EPI_ISL_601657\|20 | ···················· | ···················· | ···················· | ···················· | ···················· | ···················· | |
| N | 6283 | hCoV-19/England/MILK-9D47E9/2020\|EPI_ISL_601658\|20 | ···················· | ···················· | ···················· | ···················· | ······T············· | ···················· | |
| N | 6284 | hCoV-19/England/MILK-9D4BDE/2020\|EPI_ISL_601659\|20 | ···················· | ···················· | ···················· | ···················· | ···················· | ···················· | |
| N | 6285 | hCoV-19/Wales/MILK-9D4C71/2020\|EPI_ISL_601661\|2020 | ···················· | ···················· | ···················· | ···················· | ···················· | ···················· | |
| N | 6286 | hCoV-19/Wales/MILK-9D4C53/2020\|EPI_ISL_601662\|2020 | ···················· | ···················· | ···················· | ···················· | ···················· | ···················· | |
| N | 6287 | hCoV-19/England/CAMC-9D2314/2020\|EPI_ISL_601663\|20 | ···················· | ···················· | ···················· | ···················· | ···················· | ···················· | |
| N | 6288 | hCoV-19/England/CAMC-9D2129/2020\|EPI_ISL_601664\|20 | ···················· | ···················· | ···················· | ···················· | ···················· | ···················· | |
| N | 6289 | hCoV-19/England/CAMC-9D237E/2020\|EPI_ISL_601666\|20 | ···················· | ···················· | ···················· | ···················· | ···················· | ···················· | |
| N | 6290 | hCoV-19/England/CAMC-9D2147/2020\|EPI_ISL_601668\|20 | ···················· | ···················· | ···················· | ···················· | ···················· | ···················· | |
| N | 6291 | hCoV-19/England/CAMC-9D1EAB/2020\|EPI_ISL_601690\|20 | ···················· | ···················· | ···················· | ···················· | ······T············· | ···················· | |
| N | 6292 | hCoV-19/England/CAMC-9D229F/2020\|EPI_ISL_601691\|20 | ···················· | ·A·················· | ···················· | ···················· | ···················· | ···················· | |
| N | 6293 | hCoV-19/England/MILK-9D4B47/2020\|EPI_ISL_601692\|20 | ···················· | ···················· | ···················· | ···················· | ···················· | ···················· | |
| N | 6294 | hCoV-19/Wales/MILK-9D3BFD/2020\|EPI_ISL_601693\|2020 | ···················· | ···················· | ···················· | ···················· | ···················· | ···················· | |
| N | 6295 | hCoV-19/Wales/MILK-9D4FC3/2020\|EPI_ISL_601694\|2020 | ···················· | ···················· | ···················· | ···················· | ···················· | ···················· | |
| N | 6296 | hCoV-19/England/MILK-9D402A/2020\|EPI_ISL_601695\|20 | ···················· | ···················· | ···················· | ···················· | ···················· | ···················· | |
| N | 6297 | hCoV-19/England/MILK-9D4E5D/2020\|EPI_ISL_601699\|20 | ···················· | ···················· | ···················· | ···················· | ···················· | ···················· | |
| N | 6298 | hCoV-19/South Africa/KRISP-K003722/2020\|EPI_ISL_60 | ···················· | ·A·················· | ···················· | ···················· | ···················· | ···················· | |
| N | 6299 | hCoV-19/South Africa/KRISP-K003724/2020\|EPI_ISL_60 | ···················· | ···················· | ···················· | ···················· | ···················· | ···················· | |
| N | 6302 | hCoV-19/South Africa/KRISP-K003729/2020\|EPI_ISL_60 | ···················· | ···················· | ···················· | ···················· | ···················· | ···················· | |
| N | 6303 | hCoV-19/South Africa/KRISP-K003730/2020\|EPI_ISL_60 | ···················· | ···················· | ···················· | ···················· | ···················· | ···················· | |
| N | 6304 | hCoV-19/South Africa/KRISP-K003733/2020\|EPI_ISL_60 | ···················· | ···················· | ···················· | ···················· | ···················· | ···················· | |
| N | 6305 | hCoV-19/South Africa/KRISP-K003734/2020\|EPI_ISL_60 | ···················· | ···················· | ···················· | ···················· | ···················· | ···················· | |
| N | 6306 | hCoV-19/South Africa/KRISP-K003735/2020\|EPI_ISL_60 | ···················· | ···················· | ···················· | ···················· | ···················· | ···················· | |
| N | 6307 | hCoV-19/South Africa/KRISP-K003739/2020\|EPI_ISL_60 | ···················· | ···················· | ···················· | ···················· | ···················· | ···················· | |
| N | 6308 | hCoV-19/South Africa/KRISP-K003740/2020\|EPI_ISL_60 | ···················· | ···················· | ···················· | ···················· | ···················· | ···················· | |
| N | 6309 | hCoV-19/South Africa/KRISP-K003745/2020\|EPI_ISL_60 | ···················· | ···················· | ···················· | ···················· | ···················· | ···················· | |
| N | 6310 | hCoV-19/South Africa/KRISP-K003746/2020\|EPI_ISL_60 | ···················· | ···················· | ···················· | ···················· | ···················· | ···················· | |
| N | 6312 | hCoV-19/South Africa/KRISP-K003749/2020\|EPI_ISL_60 | ···················· | ···················· | ···················· | ···················· | ···················· | ···················· | |
| N | 6314 | hCoV-19/South Africa/KRISP-K003753/2020\|EPI_ISL_60 | ···················· | ···················· | ···················· | ···················· | ···················· | ···················· | |
| N | 6315 | hCoV-19/South Africa/KRISP-K003757/2020\|EPI_ISL_60 | ···················· | ····A··············· | ···················· | ···················· | ···················· | ···················· | |
| N | 6316 | hCoV-19/South Africa/KRISP-K003758/2020\|EPI_ISL_60 | ···················· | ···················· | ···················· | ···················· | ···················· | ···················· | |
| N | 6317 | hCoV-19/South Africa/KRISP-K003759/2020\|EPI_ISL_60 | ···················· | ···················· | ···················· | ···················· | ···················· | ···················· | |
| N | 6318 | hCoV-19/USA/MN-MDH-1843/2020\|EPI_ISL_602978\|2020-0 | ···················· | ···················· | ···················· | ···················· | ···················· | ···················· | |
| N | 6319 | hCoV-19/USA/MN-MDH-1844/2020\|EPI_ISL_602979\|2020-0 | ···················· | ···················· | ···················· | ···················· | ···················· | ···················· | |
| N | 6320 | hCoV-19/USA/MN-MDH-1845/2020\|EPI_ISL_602980\|2020-0 | ···················· | ·A·················· | ···················· | ···················· | ···················· | ···················· | |
| N | 6321 | hCoV-19/USA/MN-MDH-1846/2020\|EPI_ISL_602981\|2020-0 | ···················· | ···················· | ···················· | ···················· | ···················· | ···················· | |
| N | 6322 | hCoV-19/USA/MN-MDH-1847/2020\|EPI_ISL_602982\|2020-0 | ···················· | ·A·················· | ···················· | ···················· | ···················· | ···················· | |
| N | 6323 | hCoV-19/USA/MN-MDH-1848/2020\|EPI_ISL_602983\|2020-1 | ···················· | ···················· | ···················· | ···················· | ···················· | ···················· | |
| N | 6324 | hCoV-19/USA/MN-MDH-1849/2020\|EPI_ISL_602984\|2020-1 | ···················· | ·A·················· | ···················· | ···················· | ···················· | ···················· | |
| N | 6325 | hCoV-19/USA/MN-MDH-1850/2020\|EPI_ISL_602985\|2020-1 | ···················· | ···················· | ···················· | ···················· | ···················· | ···················· | |
| N | 6326 | hCoV-19/USA/MN-MDH-1851/2020\|EPI_ISL_602986\|2020-1 | ···················· | ·A·················· | ···················· | ···················· | ···················· | ···················· | |
| N | 6327 | hCoV-19/USA/MN-MDH-1852/2020\|EPI_ISL_602987\|2020-1 | ···················· | ·A·················· | ···················· | ···················· | ···················· | ···················· | |
| N | 6328 | hCoV-19/USA/MN-MDH-1853/2020\|EPI_ISL_602988\|2020-1 | ···················· | ·A·················· | ···················· | ···················· | ···················· | ···················· | |
| N | 6329 | hCoV-19/USA/MN-MDH-1854/2020\|EPI_ISL_602989\|2020-1 | ···················· | ·A·················· | ···················· | ···················· | ···················· | ···················· | |
| N | 6330 | hCoV-19/USA/MN-MDH-1857/2020\|EPI_ISL_602992\|2020-1 | ···················· | ···················· | ···················· | ···················· | ···················· | ···················· | |
| N | 6331 | hCoV-19/USA/MN-MDH-1858/2020\|EPI_ISL_602993\|2020-1 | ···················· | ···················· | ···················· | ···················· | ···················· | ···················· | |
| N | 6332 | hCoV-19/USA/MN-MDH-1859/2020\|EPI_ISL_602994\|2020-1 | ···················· | ···················· | ···················· | ···················· | ···················· | ···················· | |
| N | 6333 | hCoV-19/USA/MN-MDH-1861/2020\|EPI_ISL_602996\|2020-1 | ···················· | ·A·················· | ···················· | ···················· | ···················· | ···················· | |
| N | 6334 | hCoV-19/USA/MN-MDH-1862/2020\|EPI_ISL_602997\|2020-1 | ···················· | ·A·················· | ···················· | ···················· | ···················· | ···················· | |
| N | 6335 | hCoV-19/USA/MN-MDH-1863/2020\|EPI_ISL_602998\|2020-1 | ···················· | ·············A······ | ···················· | ···················· | ···················· | ···················· | |
| N | 6336 | hCoV-19/USA/MN-MDH-1864/2020\|EPI_ISL_602999\|2020-1 | ···················· | ·············A······ | ···················· | ···················· | ···················· | ···················· | |
| N | 6337 | hCoV-19/Ireland/CO-NVRL-20G32603/2020\|EPI_ISL_6051 | ···················· | ···················· | ···················· | ···················· | ···················· | ···················· | |
| N | 6338 | hCoV-19/Ireland/D-NVRL-20G35651/2020\|EPI_ISL_60513 | ···················· | ···················A | ···················· | ···················· | ···················· | ···················· | 1 |
| N | 6339 | hCoV-19/Ireland/D-NVRL-20G35116/2020\|EPI_ISL_60513 | ···················· | ···················· | ···················· | ···················· | ···················· | ···················· | |
| N | 6340 | hCoV-19/Ireland/D-NVRL-20G35434/2020\|EPI_ISL_60513 | ···················· | ···················· | ···················· | ···················· | ···················· | ···················· | |
| N | 6341 | hCoV-19/Ireland/CO-NVRL-20G32637/2020\|EPI_ISL_6051 | ···················· | ···················· | ···················· | ···················· | ···················· | ···················· | |
| N | 6342 | hCoV-19/Ireland/D-NVRL-20G32319/2020\|EPI_ISL_60513 | ···················· | ···················· | ···················· | ···················· | ···················· | ···················· | |
| N | 6343 | hCoV-19/Ireland/D-NVRL-20G32321/2020\|EPI_ISL_60513 | ···················· | ···················· | ···················· | ···················· | ···················· | ···················· | |
| N | 6344 | hCoV-19/Ireland/D-NVRL-20G35142/2020\|EPI_ISL_60514 | ···················· | ···················· | ···················· | ···················· | ···················· | ···················· | |
| N | 6345 | hCoV-19/Ireland/D-NVRL-20G32643/2020\|EPI_ISL_60514 | ···················· | ···················· | ···················· | ···················· | ···················· | ···················· | |
| N | 6346 | hCoV-19/Ireland/D-NVRL-20G35149/2020\|EPI_ISL_60514 | ···················· | ···················· | ···················· | ···················· | ···················· | ···················· | |
| N | 6347 | hCoV-19/Ireland/D-NVRL-20G32320/2020\|EPI_ISL_60514 | ···················· | ···················· | ···················· | ···················· | ···················· | ···················· | |
| N | 6348 | hCoV-19/Ireland/CO-NVRL-20G32225/2020\|EPI_ISL_6051 | ···················· | ···················· | ···················· | ···················· | ···················· | ···················· | |
| N | 6349 | hCoV-19/USA/UT-UPHL-2009383/2020\|EPI_ISL_605148\|20 | ···················· | ···················· | ···················· | ···················· | ···················· | ···················· | |
| N | 6350 | hCoV-19/USA/UT-UPHL-2009387/2020\|EPI_ISL_605150\|20 | ···················· | ···················· | ···················· | ···················· | ···················· | ···················· | |
| N | 6351 | hCoV-19/USA/UT-UPHL-2009391/2020\|EPI_ISL_605153\|20 | ···················· | ···················· | ···················· | ···················· | ···················· | ···················· | |
| N | 6352 | hCoV-19/USA/UT-UPHL-2009471/2020\|EPI_ISL_605157\|20 | ···················· | ···················· | ···················· | ···················· | ···················· | ···················· | |
| N | 6353 | hCoV-19/USA/UT-UPHL-2009472/2020\|EPI_ISL_605158\|20 | ···················· | ···················· | ···················· | ···················· | ···················· | ···················· | |

| | | | | | | | | |
|---|---|---|---|---|---|---|---|---|
| N | 6354 | hCoV-19/USA/UT-UPHL-2009473/2020\|EPI_ISL_605159\|20 | ···················· | ···················· | ···················· | ···················· | ···················· | ···················· |
| N | 6355 | hCoV-19/USA/UT-UPHL-2009475/2020\|EPI_ISL_605160\|20 | ···················· | ···················· | ···················· | ···················· | ···················· | ···················· |
| N | 6356 | hCoV-19/USA/UT-UPHL-2009478/2020\|EPI_ISL_605162\|20 | ···················· | ···················· | ···················· | ···················· | ···················· | ···················· |
| N | 6357 | hCoV-19/USA/UT-UPHL-2009480/2020\|EPI_ISL_605164\|20 | ···················· | ···················· | ···················· | ···················· | ···················· | ···················· |
| N | 6358 | hCoV-19/USA/UT-UPHL-2009481/2020\|EPI_ISL_605165\|20 | ···················· | ···················· | ···················· | ···················· | ···················· | ···················· |
| N | 6359 | hCoV-19/USA/UT-UPHL-2009482/2020\|EPI_ISL_605166\|20 | ···················· | ···················· | ···················· | ···················· | ···················· | ···················· |
| N | 6360 | hCoV-19/USA/UT-UPHL-2009486/2020\|EPI_ISL_605170\|20 | ···················· | ···················· | ···················· | ···················· | ···················· | ···················· |
| N | 6361 | hCoV-19/USA/UT-UPHL-2009488/2020\|EPI_ISL_605172\|20 | ···················· | ···················· | ···················· | ···················· | ···················· | ···················· |
| N | 6362 | hCoV-19/USA/UT-UPHL-2009489/2020\|EPI_ISL_605173\|20 | ···················· | ···················· | ···················· | ···················· | ···················· | ···················· |
| N | 6363 | hCoV-19/USA/UT-UPHL-2009493/2020\|EPI_ISL_605177\|20 | ···················· | ···················· | ···················· | ···················· | ···················· | ···················· |
| N | 6364 | hCoV-19/USA/UT-UPHL-2009497/2020\|EPI_ISL_605181\|20 | ···················· | ···················· | ···················· | ···················· | ···················· | ···················· |
| N | 6365 | hCoV-19/USA/UT-UPHL-2009498/2020\|EPI_ISL_605182\|20 | ···················· | ···················· | ···················· | ···················· | ···················· | ···················· |
| N | 6366 | hCoV-19/USA/UT-UPHL-2009499/2020\|EPI_ISL_605183\|20 | ···················· | ···················· | ···················· | ···················· | ···················· | ···················· |
| N | 6367 | hCoV-19/USA/UT-UPHL-2009503/2020\|EPI_ISL_605187\|20 | ···················· | ···················· | ···················· | ···················· | ···················· | ···················· |
| N | 6368 | hCoV-19/USA/UT-UPHL-2009509/2020\|EPI_ISL_605192\|20 | ···················· | ···················· | ···················· | ···················· | ···················· | ···················· |
| N | 6369 | hCoV-19/USA/UT-UPHL-2009510/2020\|EPI_ISL_605193\|20 | ···················· | ·A·················· | ···················· | ···················· | ···················· | ···················· |
| N | 6370 | hCoV-19/USA/UT-UPHL-2009511/2020\|EPI_ISL_605194\|20 | ···················· | ···················· | ···················· | ···················· | ···················· | ···················· |
| N | 6371 | hCoV-19/USA/UT-UPHL-2009517/2020\|EPI_ISL_605198\|20 | ···················· | ·A·················· | ···················· | ···················· | ···················· | ···················· |
| N | 6372 | hCoV-19/USA/UT-UPHL-2009519/2020\|EPI_ISL_605200\|20 | ···················· | ···················· | ···················· | ···················· | ···················· | ···················· |
| N | 6373 | hCoV-19/USA/UT-UPHL-2009521/2020\|EPI_ISL_605202\|20 | ···················· | ·A·················· | ···················· | ···················· | ···················· | ···················· |
| N | 6374 | hCoV-19/USA/UT-UPHL-2009523/2020\|EPI_ISL_605204\|20 | ···················· | ···················· | ···················· | ···················· | ···················· | ···················· |
| N | 6375 | hCoV-19/USA/UT-UPHL-2009524/2020\|EPI_ISL_605205\|20 | ···················· | ···················· | ···················· | ···················· | ···················· | ···················· |
| N | 6376 | hCoV-19/USA/UT-UPHL-2009525/2020\|EPI_ISL_605206\|20 | ···················· | ···················· | ···················· | ···················· | ···················· | ···················· |
| N | 6377 | hCoV-19/USA/UT-UPHL-2009526/2020\|EPI_ISL_605207\|20 | ···················· | ···················· | ···················· | ···················· | ···················· | ···················· |
| N | 6378 | hCoV-19/USA/UT-UPHL-2009527/2020\|EPI_ISL_605208\|20 | ···················· | ···················· | ···················· | ···················· | ···················· | ···················· |
| N | 6379 | hCoV-19/USA/UT-UPHL-2009528/2020\|EPI_ISL_605209\|20 | ···················· | ···················· | ···················· | ···················· | ···················· | ···················· |
| N | 6380 | hCoV-19/USA/UT-UPHL-2009530/2020\|EPI_ISL_605211\|20 | ···················· | ·A·················· | ···················· | ···················· | ···················· | ···················· |
| N | 6381 | hCoV-19/USA/UT-UPHL-2009534/2020\|EPI_ISL_605215\|20 | ···················· | ···················· | ···················· | ···················· | ···················· | ···················· |
| N | 6382 | hCoV-19/USA/UT-UPHL-2009536/2020\|EPI_ISL_605216\|20 | ···················· | ···················· | ···················· | ···················· | ···················· | ···················· |
| N | 6383 | hCoV-19/USA/UT-UPHL-2009537/2020\|EPI_ISL_605217\|20 | ···················· | ···················· | ···················· | ···················· | ···················· | ···················· |
| N | 6384 | hCoV-19/USA/UT-UPHL-2009538/2020\|EPI_ISL_605218\|20 | ···················· | ···················· | ···················· | ···················· | ···················· | ···················· |
| N | 6385 | hCoV-19/USA/UT-UPHL-2009541/2020\|EPI_ISL_605221\|20 | ···················· | ···················· | ···················· | ···················· | ···················· | ···················· |
| N | 6386 | hCoV-19/USA/UT-UPHL-2009543/2020\|EPI_ISL_605223\|20 | ···················· | ···················· | ···················· | ···················· | ···················· | ···················· |
| N | 6387 | hCoV-19/USA/UT-UPHL-2009545/2020\|EPI_ISL_605225\|20 | ···················· | ···················· | ···················· | ···················· | ···················· | ···················· |
| N | 6388 | hCoV-19/USA/UT-UPHL-2009546/2020\|EPI_ISL_605226\|20 | ···················· | ···················· | ···················· | ···················· | ···················· | ···················· |
| N | 6389 | hCoV-19/USA/UT-UPHL-2009551/2020\|EPI_ISL_605230\|20 | ···················· | ···················· | ···················· | ···················· | ···················· | ···················· |
| N | 6390 | hCoV-19/USA/UT-UPHL-2009553/2020\|EPI_ISL_605232\|20 | ···················· | ···················· | ···················· | ···················· | ···················· | ···················· |
| N | 6391 | hCoV-19/USA/UT-UPHL-2009557/2020\|EPI_ISL_605236\|20 | ···················· | ···················· | ···················· | ···················· | ···················· | ···················· |
| N | 6392 | hCoV-19/USA/UT-UPHL-2009559/2020\|EPI_ISL_605238\|20 | ···················· | ···················· | ···················· | ···················· | ···················· | ···················· |
| N | 6393 | hCoV-19/USA/UT-UPHL-2009560/2020\|EPI_ISL_605239\|20 | ···················· | ···················· | ···················· | ···················· | ···················· | ···················· |
| N | 6394 | hCoV-19/USA/UT-UPHL-2009561/2020\|EPI_ISL_605240\|20 | ···················· | ···················· | ···················· | ···················· | ···················· | ···················· |
| N | 6395 | hCoV-19/USA/UT-UPHL-2009563/2020\|EPI_ISL_605242\|20 | ···················· | ···················· | ···················· | ···················· | ···················· | ···················· |
| N | 6396 | hCoV-19/USA/UT-UPHL-2009564/2020\|EPI_ISL_605243\|20 | ···················· | ···················· | ···················· | ···················· | ···················· | ···················· |
| N | 6397 | hCoV-19/USA/UT-UPHL-2009565/2020\|EPI_ISL_605244\|20 | ···················· | ···················· | ···················· | ···················· | ···················· | ···················· |
| N | 6398 | hCoV-19/USA/UT-UPHL-2009566/2020\|EPI_ISL_605245\|20 | ···················· | ···················· | ···················· | ···················· | ···················· | ···················· |
| N | 6399 | hCoV-19/USA/UT-UPHL-2009568/2020\|EPI_ISL_605247\|20 | ···················· | ···················· | ···················· | ···················· | ···················· | ···················· |
| N | 6400 | hCoV-19/USA/UT-UPHL-2009570/2020\|EPI_ISL_605249\|20 | ···················· | ···················· | ···················· | ···················· | ···················· | ···················· |
| N | 6401 | hCoV-19/USA/MI-UM-10036366473/2020\|EPI_ISL_596620\| | ···················· | ·············A······ | ···················· | ···················· | ···················· | ···················· |
| N | 6402 | hCoV-19/USA/MI-UM-10036381403/2020\|EPI_ISL_596621\| | ···················· | ···················· | ···················· | ···················· | ···················· | ···················· |
| N | 6404 | hCoV-19/Ireland/D-SVUH-R9588/2020\|EPI_ISL_596628\|2 | ···················· | ···················· | ···················· | ···················· | ···················· | ···················· |
| N | 6405 | hCoV-19/Ireland/D-SVUH-R9212/2020\|EPI_ISL_596629\|2 | ···················· | ···················· | ···················· | ···················· | ···················· | ···················· |
| N | 6406 | hCoV-19/Ireland/D-SVUH-R9485/2020\|EPI_ISL_596630\|2 | ···················· | ···················· | ···················· | ···················· | ···················· | ···················· |
| N | 6407 | hCoV-19/Ireland/D-SVUH-R9592/2020\|EPI_ISL_596631\|2 | ···················· | ···················· | ···················· | ···················· | ···················· | ···················· |
| N | 6408 | hCoV-19/Ireland/D-SVUH-R9593/2020\|EPI_ISL_596632\|2 | ···················· | ···················· | ···················· | ···················· | ···················· | ···················· |
| N | 6409 | hCoV-19/Ireland/D-SVUH-R9585/2020\|EPI_ISL_596636\|2 | ···················· | ···················· | ···················· | ···················· | ···················· | ···················· |
| N | 6410 | hCoV-19/Ireland/D-SVUH-R9591/2020\|EPI_ISL_596637\|2 | ···················· | ···················· | ···················· | ···················· | ···················· | ···················· |
| N | 6411 | hCoV-19/Ireland/D-SVUH-R9218/2020\|EPI_ISL_596639\|2 | ···················· | ···················· | ···················· | ···················· | ···················· | ···················· |
| N | 6412 | hCoV-19/Ireland/D-SVUH-R9582/2020\|EPI_ISL_596640\|2 | ···················· | ···················· | ···················· | ···················· | ···················· | ···················· |
| N | 6413 | hCoV-19/Ireland/D-SVUH-R5732/2020\|EPI_ISL_596653\|2 | ···················· | ···················· | ···················· | ···················· | ···················· | ···················· |
| N | 6414 | hCoV-19/Ireland/D-SVUH-R6617/2020\|EPI_ISL_596664\|2 | ···················· | ···················· | ···················· | ···················· | ···················· | ···················· |
| N | 6415 | hCoV-19/Ireland/D-SVUH-R6766/2020\|EPI_ISL_596665\|2 | ···················· | ···················· | ···················· | ···················· | ···················· | ···················· |
| N | 6416 | hCoV-19/Ireland/D-SVUH-R6876/2020\|EPI_ISL_596666\|2 | ···················· | ···················· | ···················· | ···················· | ···················· | ···················· |
| N | 6417 | hCoV-19/Ireland/D-SVUH-R5662/2020\|EPI_ISL_596668\|2 | ···················· | ···················· | ···················· | ···················· | ···················· | ···················· |
| N | 6418 | hCoV-19/Ireland/D-SVUH-R6380/2020\|EPI_ISL_596669\|2 | ···················· | ···················· | ···················· | ···················· | ···················· | ···················· |
| N | 6419 | hCoV-19/Ireland/D-SVUH-R6456/2020\|EPI_ISL_596673\|2 | ···················· | ···················· | ···················· | ···················· | ···················· | ···················· |
| N | 6420 | hCoV-19/Ireland/D-SVUH-R6635/2020\|EPI_ISL_596674\|2 | ···················· | ···················· | ···················· | ···················· | ···················· | ···················· |
| N | 6421 | hCoV-19/Ireland/D-SVUH-R6027/2020\|EPI_ISL_596675\|2 | ···················· | ···················· | ···················· | ···················· | ···················· | ···················· |
| N | 6422 | hCoV-19/Ireland/D-SVUH-R9596/2020\|EPI_ISL_596676\|2 | ···················· | ···················· | ···················· | ···················· | ···················· | ···················· |
| N | 6423 | hCoV-19/Australia/WA114/2020\|EPI_ISL_596677\|2020-0 | ···················· | ···················· | ···················· | ···················· | ···················· | ···················· |
| N | 6424 | hCoV-19/Australia/WA115/2020\|EPI_ISL_596678\|2020-0 | ···················· | ···················· | ···················· | ···················· | ···················· | ···················· |
| N | 6425 | hCoV-19/Australia/WA116/2020\|EPI_ISL_596679\|2020-0 | ···················· | ···················· | ···················· | ···················· | ···················· | ···················· |
| N | 6426 | hCoV-19/Australia/WA117/2020\|EPI_ISL_596680\|2020-0 | ···················· | ···················· | ···················· | ···················· | ···················· | ···················· |
| N | 6427 | hCoV-19/Australia/WA118/2020\|EPI_ISL_596681\|2020-0 | ···················· | ···················· | ···················· | ···················· | ···················· | ···················· |
| N | 6428 | hCoV-19/Australia/WA119/2020\|EPI_ISL_596682\|2020-0 | ···················· | ···················· | ···················· | ···················· | ···················· | ···················· |
| N | 6429 | hCoV-19/Australia/WA120/2020\|EPI_ISL_596683\|2020-0 | ···················· | ···················· | ···················· | ···················· | ···················· | ···················· |
| N | 6430 | hCoV-19/Australia/WA121/2020\|EPI_ISL_596684\|2020-0 | ···················· | ···················· | ···················· | ···················· | ···················· | ···················· |
| N | 6431 | hCoV-19/Australia/WA122/2020\|EPI_ISL_596685\|2020-0 | ···················· | ···················· | ···················· | ···················· | ···················· | ···················· |
| N | 6432 | hCoV-19/Australia/WA123/2020\|EPI_ISL_596686\|2020-0 | ···················· | ···················· | ···················· | ···················· | ···················· | ···················· |
| N | 6433 | hCoV-19/Australia/WA124/2020\|EPI_ISL_596687\|2020-0 | ···················· | ···················· | ···················· | ···················· | ···················· | ···················· |
| N | 6434 | hCoV-19/Australia/WA125/2020\|EPI_ISL_596688\|2020-0 | ···················· | ···················· | ···················· | ···················· | ···················· | ···················· |

| | | | | | | | | |
|---|---|---|---|---|---|---|---|---|
| N | 6435 | hCoV-19/Australia/WA126/2020\|EPI_ISL_596689\|2020-0 | .................... | .................... | ..................... | .................... | .................... | .................... |
| N | 6436 | hCoV-19/Australia/WA127/2020\|EPI_ISL_596690\|2020-0 | .................... | .................... | ..................... | .................... | .................... | .................... |
| N | 6437 | hCoV-19/Australia/WA128/2020\|EPI_ISL_596691\|2020-0 | .................... | .................... | ..................... | .................... | .................... | .................... |
| N | 6438 | hCoV-19/Australia/WA129/2020\|EPI_ISL_596692\|2020-0 | .................... | .................... | ..................... | .................... | .................... | .................... |
| N | 6439 | hCoV-19/Australia/WA130/2020\|EPI_ISL_596693\|2020-0 | .................... | .................... | ..................... | .................... | .................... | .................... |
| N | 6440 | hCoV-19/Australia/WA131/2020\|EPI_ISL_596694\|2020-0 | .................... | .................... | ..................... | .................... | .................... | .................... |
| N | 6441 | hCoV-19/Australia/WA132/2020\|EPI_ISL_596695\|2020-0 | .................... | .................... | ..................... | .................... | .................... | .................... |
| N | 6442 | hCoV-19/Australia/WA133/2020\|EPI_ISL_596696\|2020-0 | .................... | .................... | ..................... | .................... | .................... | .................... |
| N | 6443 | hCoV-19/Australia/WA134/2020\|EPI_ISL_596697\|2020-0 | .................... | .................... | ..................... | .................... | .................... | .................... |
| N | 6444 | hCoV-19/Australia/WA135/2020\|EPI_ISL_596698\|2020-0 | .................... | .................... | ..................... | .................... | .................... | .................... |
| N | 6445 | hCoV-19/Australia/WA136/2020\|EPI_ISL_596699\|2020-0 | .................... | .A.................. | ..................... | .................... | .................... | ..A................. |
| N | 6446 | hCoV-19/England/ALDP-A50F19/2020\|EPI_ISL_597810\|20 | .................... | .................... | ..................... | .................... | .................... | .................... |
| N | 6447 | hCoV-19/England/ALDP-A4FBBF/2020\|EPI_ISL_597813\|20 | .................... | .................... | ..................... | .................... | .................... | .................... |
| N | 6448 | hCoV-19/England/ALDP-A50B15/2020\|EPI_ISL_597814\|20 | .................... | .................... | ..................... | .................... | .................... | .................... |
| N | 6449 | hCoV-19/England/ALDP-A5077B/2020\|EPI_ISL_597815\|20 | .................... | .................... | ..................... | .................... | .................... | .................... |
| N | 6450 | hCoV-19/England/ALDP-A4FEC5/2020\|EPI_ISL_597816\|20 | .................... | .................... | ..................... | .................... | .................... | .................... |
| N | 6451 | hCoV-19/England/ALDP-A50EB2/2020\|EPI_ISL_597817\|20 | .................... | .................... | ..................... | .................... | .................... | .................... |
| N | 6452 | hCoV-19/England/ALDP-A4FE5C/2020\|EPI_ISL_597818\|20 | .................... | .................... | ..................... | .................... | .................... | .................... |
| N | 6453 | hCoV-19/England/ALDP-A4FB0A/2020\|EPI_ISL_597819\|20 | .................... | .................... | ..................... | .................... | .................... | .................... |
| N | 6454 | hCoV-19/England/ALDP-A5008F/2020\|EPI_ISL_597827\|20 | .................... | .................... | ..................... | .................... | .................... | .................... |
| N | 6455 | hCoV-19/England/ALDP-A502D4/2020\|EPI_ISL_597829\|20 | .................... | .................... | ..................... | .................... | .................... | .................... |
| N | 6456 | hCoV-19/England/ALDP-A51367/2020\|EPI_ISL_597951\|20 | .................... | .................... | ..................... | .................... | .................... | .................... |
| N | 6457 | hCoV-19/England/ALDP-A52436/2020\|EPI_ISL_597952\|20 | .................... | .................... | ..................... | .................... | .................... | .................... |
| N | 6458 | hCoV-19/England/ALDP-A5231B/2020\|EPI_ISL_597954\|20 | .................... | .................... | ..................... | .................... | .................... | .................... |
| N | 6459 | hCoV-19/England/ALDP-A517D4/2020\|EPI_ISL_597955\|20 | .................... | .................... | ..................... | .................... | .................... | .................... |
| N | 6460 | hCoV-19/England/ALDP-A5125B/2020\|EPI_ISL_597956\|20 | .................... | .................... | ..................... | .................... | .................... | .................... |
| N | 6461 | hCoV-19/England/ALDP-A527B5/2020\|EPI_ISL_597957\|20 | .................... | .................... | ..................... | .................... | .................... | .................... |
| N | 6462 | hCoV-19/England/ALDP-A5181D/2020\|EPI_ISL_597958\|20 | .................... | .................... | ..................... | .................... | .................... | .................... |
| N | 6463 | hCoV-19/England/ALDP-A5282B/2020\|EPI_ISL_597959\|20 | .................... | .................... | ..................... | .................... | .................... | .................... |
| N | 6464 | hCoV-19/England/ALDP-A520C9/2020\|EPI_ISL_597960\|20 | .................... | .................... | ..................... | .................... | .................... | .................... |
| N | 6465 | hCoV-19/England/ALDP-A5133A/2020\|EPI_ISL_597961\|20 | .................... | .................... | ..................... | .................... | .................... | .................... |
| N | 6466 | hCoV-19/England/ALDP-A5215D/2020\|EPI_ISL_597962\|20 | .................... | .................... | ..................... | .................... | .................... | .................... |
| N | 6467 | hCoV-19/England/ALDP-A51D0F/2020\|EPI_ISL_597963\|20 | .................... | .................... | ..................... | .................... | .................... | .................... |
| N | 6468 | hCoV-19/England/ALDP-A5166E/2020\|EPI_ISL_597965\|20 | .................... | .................... | ..................... | .................... | .................... | .................... |
| N | 6469 | hCoV-19/England/ALDP-A5130D/2020\|EPI_ISL_597966\|20 | .................... | .................... | ..................... | .................... | .................... | .................... |
| N | 6470 | hCoV-19/England/ALDP-A525AC/2020\|EPI_ISL_597967\|20 | .................... | .................... | ..................... | .................... | .................... | .................... |
| N | 6471 | hCoV-19/England/ALDP-A513D0/2020\|EPI_ISL_597968\|20 | .................... | .................... | ..................... | .................... | .................... | .................... |
| N | 6472 | hCoV-19/England/ALDP-A52199/2020\|EPI_ISL_597969\|20 | .................... | .................... | ..................... | .................... | .................... | .................... |
| N | 6473 | hCoV-19/England/ALDP-A522C3/2020\|EPI_ISL_597970\|20 | .................... | .................... | ..................... | .................... | .................... | .................... |
| N | 6474 | hCoV-19/England/ALDP-A51B6F/2020\|EPI_ISL_597971\|20 | .................... | .................... | ..................... | .................... | .................... | .................... |
| N | 6475 | hCoV-19/England/ALDP-A514CE/2020\|EPI_ISL_597972\|20 | .................... | .................... | ..................... | .................... | .................... | .................... |
| N | 6476 | hCoV-19/England/ALDP-A52542/2020\|EPI_ISL_597985\|20 | .................... | .................... | ..................... | .................... | .................... | .................... |
| N | 6477 | hCoV-19/England/ALDP-A5271F/2020\|EPI_ISL_597986\|20 | .................... | .................... | ..................... | .................... | .................... | .................... |
| N | 6478 | hCoV-19/England/ALDP-A51D3C/2020\|EPI_ISL_597987\|20 | .................... | .................... | ..................... | .................... | .................... | .................... |
| N | 6479 | hCoV-19/England/ALDP-A520AB/2020\|EPI_ISL_597988\|20 | .................... | .................... | ..................... | .................... | .................... | .................... |
| N | 6480 | hCoV-19/England/ALDP-A51CF3/2020\|EPI_ISL_597989\|20 | .................... | .................... | ..................... | .................... | .................... | .................... |
| N | 6481 | hCoV-19/England/ALDP-A51859/2020\|EPI_ISL_597995\|20 | .................... | .................... | ..................... | .................... | .................... | .................... |
| N | 6482 | hCoV-19/England/ALDP-A52339/2020\|EPI_ISL_597996\|20 | .................... | .................... | ..................... | .................... | .................... | .................... |
| N | 6483 | hCoV-19/England/ALDP-A51EDF/2020\|EPI_ISL_597997\|20 | .................... | .................... | ..................... | .................... | .................... | .................... |
| N | 6484 | hCoV-19/England/ALDP-A51E1B/2020\|EPI_ISL_597998\|20 | .................... | .................... | ..................... | .................... | .................... | .................... |
| N | 6485 | hCoV-19/England/ALDP-A51CD5/2020\|EPI_ISL_597999\|20 | .................... | .................... | ..................... | .................... | .................... | .................... |
| N | 6486 | hCoV-19/Scotland/QEUH-A0069B/2020\|EPI_ISL_600239\|2 | .................... | .................... | ..................... | .................... | .................... | .................... |
| N | 6487 | hCoV-19/England/QEUH-A0015E/2020\|EPI_ISL_600240\|20 | .................... | .................... | ..................... | .................... | .................... | .................... |
| N | 6488 | hCoV-19/England/QEUH-9FF9A6/2020\|EPI_ISL_600241\|20 | .................... | .................... | ..................... | .................... | .................... | .................... |
| N | 6489 | hCoV-19/England/QEUH-A002B5/2020\|EPI_ISL_600242\|20 | .................... | .................... | ..................... | .................... | .................... | .................... |
| N | 6490 | hCoV-19/England/QEUH-9FFF59/2020\|EPI_ISL_600243\|20 | .................... | .................... | ..................... | .................... | .................... | .................... |
| N | 6491 | hCoV-19/Scotland/QEUH-9FF7BB/2020\|EPI_ISL_600244\|2 | .................... | .................... | ..................... | .................... | .................... | .................... |
| N | 6492 | hCoV-19/Scotland/QEUH-9FFF0E/2020\|EPI_ISL_600245\|2 | .................... | .................... | ..................... | .................... | .................... | .................... |
| N | 6493 | hCoV-19/Scotland/QEUH-9FFEC5/2020\|EPI_ISL_600246\|2 | .................... | .................... | ..................... | .................... | .................... | .................... |
| N | 6494 | hCoV-19/England/QEUH-A0009D/2020\|EPI_ISL_600247\|20 | .................... | .................... | ..................... | .................... | .................... | .................... |
| N | 6495 | hCoV-19/England/QEUH-9FFA2B/2020\|EPI_ISL_600248\|20 | .................... | .................... | ..................... | .................... | .................... | .................... |
| N | 6496 | hCoV-19/England/QEUH-A0081D/2020\|EPI_ISL_600249\|20 | .................... | .................... | ..................... | .................... | .................... | .................... |
| N | 6497 | hCoV-19/England/QEUH-A0021F/2020\|EPI_ISL_600254\|20 | .................... | .................... | ..................... | .................... | .................... | .................... |
| N | 6498 | hCoV-19/England/QEUH-A00358/2020\|EPI_ISL_600255\|20 | .................... | .................... | ..................... | .................... | .................... | .................... |
| N | 6499 | hCoV-19/England/QEUH-9FF539/2020\|EPI_ISL_600256\|20 | .................... | .A.................. | ..................... | .................... | .................... | .................... |
| N | 6500 | hCoV-19/England/QEUH-9FFD5F/2020\|EPI_ISL_600280\|20 | .................... | .................... | ..................... | .................... | .................... | .................... |
| N | 6501 | hCoV-19/England/QEUH-9FFB19/2020\|EPI_ISL_600281\|20 | .................... | .................... | ..................... | .................... | .................... | .................... |
| N | 6502 | hCoV-19/England/QEUH-9FF171/2020\|EPI_ISL_600282\|20 | .................... | .................... | ..................... | .................... | .................... | .................... |
| N | 6503 | hCoV-19/England/QEUH-A00631/2020\|EPI_ISL_600283\|20 | .................... | .................... | ..................... | .................... | .................... | .................... |
| N | 6504 | hCoV-19/England/QEUH-9FF6EB/2020\|EPI_ISL_600284\|20 | .................... | .................... | ..................... | .................... | .................... | .................... |
| N | 6505 | hCoV-19/England/QEUH-9FFF86/2020\|EPI_ISL_600285\|20 | .................... | .................... | ..................... | .................... | .................... | .................... |
| N | 6506 | hCoV-19/England/QEUH-9FF42D/2020\|EPI_ISL_600286\|20 | .................... | .................... | ..................... | .................... | .................... | .................... |
| N | 6507 | hCoV-19/England/QEUH-9FFF4A/2020\|EPI_ISL_600287\|20 | .................... | .................... | ..................... | .................... | .................... | .................... |
| N | 6508 | hCoV-19/England/QEUH-A0031C/2020\|EPI_ISL_600288\|20 | .................... | .................... | ..................... | .................... | .................... | .................... |
| N | 6509 | hCoV-19/Scotland/QEUH-A00789/2020\|EPI_ISL_600289\|2 | .................... | .................... | ..................... | .................... | .................... | .................... |
| N | 6510 | hCoV-19/England/QEUH-9FF93D/2020\|EPI_ISL_600290\|20 | .................... | .................... | ..................... | .................... | .................... | .................... |
| N | 6511 | hCoV-19/England/QEUH-9FF2BA/2020\|EPI_ISL_600291\|20 | .................... | .................... | ..................... | .................... | .................... | .................... |
| N | 6512 | hCoV-19/England/QEUH-A0072F/2020\|EPI_ISL_600292\|20 | .................... | .................... | ..................... | .................... | .................... | .................... |
| N | 6513 | hCoV-19/England/QEUH-A004CE/2020\|EPI_ISL_600293\|20 | .................... | .................... | ..................... | .................... | .................... | .................... |
| N | 6514 | hCoV-19/England/QEUH-9FF654/2020\|EPI_ISL_600294\|20 | .................... | .................... | ..................... | .................... | .................... | .................... |

| | | | | | | | | |
|---|---|---|---|---|---|---|---|---|
| N | 6515 | hCoV-19/England/QEUH-9FFD6E/2020\|EPI_ISL_600295\|20 | ···················· | ·A·················· | ···················· | ···················· | ···················· | ···················· |
| N | 6516 | hCoV-19/Scotland/QEUH-9FFED4/2020\|EPI_ISL_600296\|2 | ···················· | ···················· | ···················· | ···················· | ···················· | ···················· |
| N | 6517 | hCoV-19/England/QEUH-A00974/2020\|EPI_ISL_600298\|20 | ···················· | ···················· | ···················· | ···················· | ···················· | ···················· |
| N | 6518 | hCoV-19/England/QEUH-9FF44B/2020\|EPI_ISL_600299\|20 | ···················· | ···················· | ···················· | ···················· | ···················· | ···················· |
| N | 6519 | hCoV-19/Scotland/QEUH-9FF7F7/2020\|EPI_ISL_600300\|2 | ···················· | ···················· | ···················· | ···················· | ···················· | ···················· |
| N | 6520 | hCoV-19/England/QEUH-9FF1DB/2020\|EPI_ISL_600301\|20 | ···················· | ···················· | ···················· | ···················· | ···················· | ···················· |
| N | 6521 | hCoV-19/England/QEUH-9FF43C/2020\|EPI_ISL_600303\|20 | ···················· | ···················· | ···················· | ···················· | ······T············· | ···················· |
| N | 6522 | hCoV-19/England/QEUH-9FF8B8/2020\|EPI_ISL_600304\|20 | ···················· | ···················· | ···················· | ···················· | ···················· | ···················· |
| N | 6523 | hCoV-19/England/QEUH-9FF2AB/2020\|EPI_ISL_600305\|20 | ···················· | ···················· | ···················· | ···················· | ···················· | ···················· |
| N | 6524 | hCoV-19/England/QEUH-9FF29C/2020\|EPI_ISL_600307\|20 | ···················· | ···················· | ···················· | ···················· | ···················· | ···················· |
| N | 6525 | hCoV-19/England/QEUH-A00288/2020\|EPI_ISL_600309\|20 | ···················· | ···················· | ···················· | ···················· | ···················· | ···················· |
| N | 6526 | hCoV-19/England/QEUH-A002E2/2020\|EPI_ISL_600310\|20 | ···················· | ·A·················· | ···················· | ···················· | ···················· | ···················· |
| N | 6527 | hCoV-19/England/QEUH-A0023D/2020\|EPI_ISL_600311\|20 | ···················· | ···················· | ···················· | ···················· | ···················· | ···················· |
| N | 6528 | hCoV-19/England/QEUH-9FF302/2020\|EPI_ISL_600312\|20 | ···················· | ···················· | ···················· | ···················· | ···················· | ···················· |
| N | 6529 | hCoV-19/England/QEUH-9FFDC8/2020\|EPI_ISL_600313\|20 | ···················· | ···················· | ···················· | ···················· | ···················· | ···················· |
| N | 6530 | hCoV-19/England/QEUH-A0018B/2020\|EPI_ISL_600314\|20 | ···················· | ···················· | ···················· | ···················· | ···················· | ···················· |
| N | 6531 | hCoV-19/England/QEUH-A0073E/2020\|EPI_ISL_600315\|20 | ···················· | ···················· | ···················· | ···················· | ···················· | ···················· |
| N | 6532 | hCoV-19/England/QEUH-9FFC52/2020\|EPI_ISL_600334\|20 | ···················· | ···················· | ···················· | ···················· | ···················· | ···················· |
| N | 6533 | hCoV-19/England/QEUH-9FFF68/2020\|EPI_ISL_600338\|20 | ···················· | ···················· | ···················· | ···················· | ···················· | ···················· |
| N | 6534 | hCoV-19/Scotland/QEUH-A003FE/2020\|EPI_ISL_600339\|2 | ···················· | ···················· | ···················· | ···················· | ···················· | ···················· |
| N | 6535 | hCoV-19/England/QEUH-A000E8/2020\|EPI_ISL_600340\|20 | ···················· | ···················· | ···················· | ···················· | ···················· | ···················· |
| N | 6536 | hCoV-19/England/QEUH-9FF618/2020\|EPI_ISL_600342\|20 | ···················· | ···················· | ···················· | ···················· | ···················· | ···················· |
| N | 6537 | hCoV-19/England/QEUH-A000BB/2020\|EPI_ISL_600343\|20 | ···················· | ···················· | ···················· | ···················· | ···················· | ···················· |
| N | 6538 | hCoV-19/England/QEUH-9FFE01/2020\|EPI_ISL_600344\|20 | ···················· | ···················· | ···················· | ···················· | ···················· | ···················· |
| N | 6539 | hCoV-19/England/QEUH-9FF51B/2020\|EPI_ISL_600345\|20 | ···················· | ···················· | ···················· | ···················· | ···················· | ···················· |
| N | 6540 | hCoV-19/England/QEUH-9FF733/2020\|EPI_ISL_600347\|20 | ···················· | ···················· | ···················· | ···················· | ···················· | ···················· |
| N | 6541 | hCoV-19/England/QEUH-9FF162/2020\|EPI_ISL_600348\|20 | ···················· | ···················· | ···················· | ···················· | ···················· | ···················· |
| N | 6542 | hCoV-19/England/QEUH-9FF2D8/2020\|EPI_ISL_600349\|20 | ···················· | ···················· | ···················· | ···················· | ···················· | ···················· |
| N | 6543 | hCoV-19/England/QEUH-A000D9/2020\|EPI_ISL_600350\|20 | ···················· | ···················· | ···················· | ···················· | ···················· | ···················· |
| N | 6544 | hCoV-19/Scotland/QEUH-9FF77F/2020\|EPI_ISL_600351\|2 | ···················· | ···················· | ···················· | ···················· | ···················· | ···················· |
| N | 6545 | hCoV-19/England/QEUH-9FF92E/2020\|EPI_ISL_600352\|20 | ···················· | ···················· | ···················· | ···················· | ···················· | ···················· |
| N | 6546 | hCoV-19/Scotland/QEUH-9FFE6B/2020\|EPI_ISL_600353\|2 | ···················· | ···················· | ···················· | ···················· | ···················· | ···················· |
| N | 6547 | hCoV-19/England/QEUH-A00024/2020\|EPI_ISL_600354\|20 | ···················· | ···················· | ···················· | ···················· | ···················· | ···················· |
| N | 6548 | hCoV-19/England/QEUH-9FF724/2020\|EPI_ISL_600355\|20 | ···················· | ···················· | ···················· | ···················· | ······T············· | ···················· |
| N | 6549 | hCoV-19/England/QEUH-9FFF77/2020\|EPI_ISL_600356\|20 | ···················· | ···················· | ···················· | ···················· | ···················· | ···················· |
| N | 6550 | hCoV-19/England/QEUH-9FF4D2/2020\|EPI_ISL_600357\|20 | ···················· | ···················· | ···················· | ···················· | ···················· | ···················· |
| N | 6551 | hCoV-19/England/QEUH-9FFD40/2020\|EPI_ISL_600358\|20 | ···················· | ···················· | ···················· | ···················· | ···················· | ···················· |
| N | 6552 | hCoV-19/England/QEUH-A0019A/2020\|EPI_ISL_600359\|20 | ···················· | ···················· | ···················· | ···················· | ···················· | ···················· |
| N | 6553 | hCoV-19/England/MILK-9D4A77/2020\|EPI_ISL_601570\|20 | ···················· | ···················· | ···················· | ···················· | ···················· | ···················· |
| N | 6554 | hCoV-19/England/CAMC-9D1E7E/2020\|EPI_ISL_601571\|20 | ···················· | ···················· | ···················· | ···················· | ···················· | ···················· |
| N | 6555 | hCoV-19/England/MILK-9D40FD/2020\|EPI_ISL_601572\|20 | ···················· | ···················· | ···················· | ···················· | ···················· | ···················· |
| N | 6556 | hCoV-19/England/MILK-9D4A2C/2020\|EPI_ISL_601574\|20 | ···················· | ···················· | ···················· | ···················· | ···················· | ···················· |
| N | 6557 | hCoV-19/England/MILK-9D4B56/2020\|EPI_ISL_601575\|20 | ···················· | ···················· | ···················· | ···················· | ···················· | ···················· |
| N | 6558 | hCoV-19/England/MILK-9D4048/2020\|EPI_ISL_601576\|20 | ···················· | ···················· | ···················· | ···················· | ···················· | ···················· |
| N | 6559 | hCoV-19/England/MILK-9D4FF0/2020\|EPI_ISL_601577\|20 | ···················· | ···················· | ···················· | ···················· | ···················· | ···················· |
| N | 6560 | hCoV-19/England/CAMC-9D2323/2020\|EPI_ISL_601578\|20 | ···················· | ···················· | ···················· | ···················· | ···················· | ···················· |
| N | 6561 | hCoV-19/England/CAMC-9D236F/2020\|EPI_ISL_601579\|20 | ···················· | ···················· | ···················· | ···················· | ···················· | ···················· |
| N | 6562 | hCoV-19/England/CAMC-9D1E23/2020\|EPI_ISL_601585\|20 | ···················· | ···················· | ···················· | ···················· | ···················· | ···················· |
| N | 6563 | hCoV-19/England/MILK-9D4A95/2020\|EPI_ISL_601586\|20 | ···················· | ···················· | ···················· | ···················· | ···················· | ···················· |
| N | 6564 | hCoV-19/England/MILK-9D4DD8/2020\|EPI_ISL_601587\|20 | ···················· | ···················· | ···················· | ···················· | ···················· | ···················· |
| N | 6565 | hCoV-19/England/MILK-9D4998/2020\|EPI_ISL_601603\|20 | ···················· | ···················· | ···················· | ···················· | ···················· | ···················· |
| N | 6566 | hCoV-19/England/CAMC-9D23C9/2020\|EPI_ISL_601604\|20 | ···················· | ···················· | ···················· | ···················· | ···················· | ···················· |
| N | 6567 | hCoV-19/England/MILK-9D4084/2020\|EPI_ISL_601605\|20 | ···················· | ···················· | ···················· | ···················· | ···················· | ···················· |
| N | 6568 | hCoV-19/England/CAMC-9D1EF6/2020\|EPI_ISL_601606\|20 | ···················· | ···················· | ···················· | ···················· | ···················· | ···················· |
| N | 6569 | hCoV-19/Wales/MILK-9D4C62/2020\|EPI_ISL_601607\|2020 | ···················· | ···················· | ···················· | ···················· | ···················· | ···················· |
| N | 6570 | hCoV-19/England/MILK-9D4145/2020\|EPI_ISL_601608\|20 | ···················· | ···················· | ···················· | ···················· | ···················· | ···················· |
| N | 6571 | hCoV-19/Wales/MILK-9D4CAE/2020\|EPI_ISL_601609\|2020 | ···················· | ···················· | ···················· | ···················· | ···················· | ···················· |
| N | 6572 | hCoV-19/England/MILK-9D4B83/2020\|EPI_ISL_601610\|20 | ···················· | ···················· | ···················· | ···················· | ···················· | ···················· |
| N | 6573 | hCoV-19/England/CAMC-9D2095/2020\|EPI_ISL_601611\|20 | ···················· | ·A·················· | ···················· | ···················· | ···················· | ···················· |
| N | 6574 | hCoV-19/England/MILK-9D3D33/2020\|EPI_ISL_601612\|20 | ···················· | ···················· | ···················· | ···················· | ···················· | ···················· |
| N | 6575 | hCoV-19/Wales/MILK-9D4CCC/2020\|EPI_ISL_601613\|2020 | ···················· | ···················· | ···················· | ···················· | ···················· | ···················· |
| N | 6576 | hCoV-19/England/MILK-9D50FC/2020\|EPI_ISL_601614\|20 | ···················· | ···················· | ···················· | ···················· | ···················· | ···················· |
| N | 6577 | hCoV-19/England/CAMC-9D1D53/2020\|EPI_ISL_601615\|20 | ···················· | ···················· | ···················· | ···················· | ···················· | ···················· |
| N | 6578 | hCoV-19/England/CAMC-9D20B3/2020\|EPI_ISL_601617\|20 | ···················· | ···················· | ···················· | ···················· | ···················· | ···················· |
| N | 6579 | hCoV-19/England/CAMC-9D1E41/2020\|EPI_ISL_601618\|20 | ···················· | ···················· | ···················· | ···················· | ···················· | ···················· |
| N | 6580 | hCoV-19/England/CAMC-9D21A1/2020\|EPI_ISL_601619\|20 | ···················· | ···················· | ···················· | ···················· | ···················· | ···················· |
| N | 6581 | hCoV-19/Wales/MILK-9D4CBD/2020\|EPI_ISL_601620\|2020 | ···················· | ···················· | ···················· | ···················· | ···················· | ···················· |
| N | 6582 | hCoV-19/Wales/MILK-9D3A2D/2020\|EPI_ISL_601621\|2020 | ···················· | ···················· | ···················· | ···················· | ···················· | ···················· |
| N | 6583 | hCoV-19/England/MILK-9D40C0/2020\|EPI_ISL_601623\|20 | ···················· | ···················· | ········T··········· | ···················· | ······T············· | ···················· |
| N | 6584 | hCoV-19/Wales/MILK-9D4497/2020\|EPI_ISL_601625\|2020 | ···················· | ···················· | ···················· | ···················· | ···················· | ···················· |
| N | 6585 | hCoV-19/England/MILK-9D4093/2020\|EPI_ISL_601626\|20 | ···················· | ·A·················· | ···················· | ···················· | ···················· | ···A················ |
| N | 6586 | hCoV-19/England/CAMC-9D1F8A/2020\|EPI_ISL_601627\|20 | ···················· | ···················· | ···················· | ···················· | ···················· | ···················· |
| N | 6587 | hCoV-19/England/CAMC-9D2226/2020\|EPI_ISL_601628\|20 | ···················· | ···················· | ···················· | ···················· | ···················· | ···················· |
| N | 6588 | hCoV-19/England/MILK-9D40B1/2020\|EPI_ISL_601629\|20 | ···················· | ···················· | ···················· | ···················· | ···················· | ···················· |
| N | 6589 | hCoV-19/England/MILK-9D3E21/2020\|EPI_ISL_601630\|20 | ···················· | ·A·················· | ···················· | ···················· | ···················· | ···················· |
| N | 6590 | hCoV-19/England/MILK-9D47BC/2020\|EPI_ISL_601632\|20 | ···················· | ···················· | ···················· | ···················· | ···················· | ···················· |
| N | 6591 | hCoV-19/England/CAMC-9D20A4/2020\|EPI_ISL_601634\|20 | ···················· | ···················· | ···················· | ···················· | ···················· | ···················· |
| N | 6592 | hCoV-19/England/MILK-9D3C54/2020\|EPI_ISL_601635\|20 | ···················· | ·A·················· | ···················· | ···················· | ···················· | ···················· |
| N | 6593 | hCoV-19/Wales/MILK-9D3C27/2020\|EPI_ISL_601637\|2020 | ···················· | ···················· | ···················· | ···················· | ···················· | ···················· |
| N | 6594 | hCoV-19/Wales/MILK-9D4619/2020\|EPI_ISL_601638\|2020 | ···················· | ···················· | ···················· | ···················· | ···················· | ···················· |

```
N    6595 hCoV-19/England/CAMC-9D1E6F/2020|EPI_ISL_601639|20    ····················  ····················  ·····················  ····················  ····················  ····················
N    6596 hCoV-19/England/MILK-9D3E9A/2020|EPI_ISL_601640|20    ····················  ····················  ·····················  ····················  ····················  ····················
N    6597 hCoV-19/England/MILK-9D4567/2020|EPI_ISL_601642|20    ····················  ····················  ·····················  ····················  ····T···············  ····················
N    6598 hCoV-19/Wales/MILK-9D4D9C/2020|EPI_ISL_601643|2020    ····················  ····················  ·····················  ····················  ····················  ····················
N    6599 hCoV-19/England/MILK-9D4901/2020|EPI_ISL_601644|20    ····················  ·A··················  ·····················  ····················  ····················  ···A················
N    6600 hCoV-19/England/MILK-9D3E30/2020|EPI_ISL_601645|20    ····················  ····················  ·····················  ····················  ····················  ····················
N    6601 hCoV-19/Wales/MILK-9D3BC0/2020|EPI_ISL_601646|2020    ····················  ····················  ·····················  ····················  ····················  ····················
N    6602 hCoV-19/England/MILK-9D4AFF/2020|EPI_ISL_601660|20    ····················  C···················  ·····················  ····················  ····················  ····················
N    6603 hCoV-19/Wales/MILK-9D450D/2020|EPI_ISL_601665|2020    ····················  ····················  ·····················  ····················  ····················  ····················
N    6604 hCoV-19/England/MILK-9D451C/2020|EPI_ISL_601669|20    ····················  ·A··················  ·····················  ····················  ····················  ···A················
N    6605 hCoV-19/England/MILK-9D4D6F/2020|EPI_ISL_601670|20    ····················  ····················  ·····················  ····················  ····················  ····················
N    6606 hCoV-19/England/MILK-9D44F1/2020|EPI_ISL_601672|20    ····················  ····················  ·····················  ····················  ····················  ····················
N    6607 hCoV-19/England/CAMC-9D21DE/2020|EPI_ISL_601673|20    ····················  ·A··················  ·····················  ····················  ····················  ···A················
N    6608 hCoV-19/England/MILK-9D453A/2020|EPI_ISL_601674|20    ····················  ····················  ·····················  ····················  ····················  ····················
N    6609 hCoV-19/Wales/MILK-9D4DE7/2020|EPI_ISL_601675|2020    ····················  ····················  ·····················  ····················  ····················  ····················
N    6610 hCoV-19/England/MILK-9D4F69/2020|EPI_ISL_601676|20    ····················  C···················  ·····················  ····················  ····················  ····················
N    6611 hCoV-19/England/MILK-9D46CE/2020|EPI_ISL_601677|20    ····················  ····················  ·····················  ····················  ····················  ····················
N    6612 hCoV-19/England/MILK-9D3B39/2020|EPI_ISL_601678|20    ····················  ····················  ·····················  ····················  ····················  ····················
N    6613 hCoV-19/Wales/MILK-9D4DAB/2020|EPI_ISL_601679|2020    ····················  ····················  ·····················  ····················  ····················  ····················
N    6614 hCoV-19/Wales/MILK-9D3A78/2020|EPI_ISL_601681|2020    ····················  ····················  ·····················  ····················  ····················  ····················
N    6615 hCoV-19/England/MILK-9D3FE2/2020|EPI_ISL_601683|20    ····················  ····················  ·····················  ····················  ····················  ····················
N    6616 hCoV-19/Wales/MILK-9D3FB5/2020|EPI_ISL_601684|2020    ····················  ····················  ·····················  ····················  ····················  ····················
N    6617 hCoV-19/England/CAMC-9D22F9/2020|EPI_ISL_601685|20    ····················  ····················  ·····················  ····················  ····················  ····················
N    6618 hCoV-19/England/MILK-9D3C63/2020|EPI_ISL_601686|20    ····················  ····················  ·····················  ····················  ····················  ····················
N    6619 hCoV-19/Wales/MILK-9D4D14/2020|EPI_ISL_601687|2020    ····················  ····················  ·····················  ····················  ····················  ····················
N    6620 hCoV-19/England/MILK-9D4664/2020|EPI_ISL_601688|20    ····················  ····················  ·····················  ····················  ······T·············  ····················
N    6621 hCoV-19/England/MILK-9D3FD3/2020|EPI_ISL_601689|20    ····················  ····················  ·····················  ····················  ····················  ····················
N    6622 hCoV-19/USA/UT-UPHL-201032745/2020|EPI_ISL_602934|    ····················  ····················  ·····················  ····················  ····················  ····················
N    6623 hCoV-19/USA/UT-UPHL-201027619/2020|EPI_ISL_602936|    ····················  ····················  ·····················  ····················  ····················  ····················
N    6624 hCoV-19/USA/UT-UPHL-201001416/2020|EPI_ISL_602946|    ····················  ····················  ·····················  ····················  ····················  ····················
N    6625 hCoV-19/USA/UT-UPHL-201012054/2020|EPI_ISL_602947|    ····················  ····················  ·····················  ····················  ····················  ····················
N    6626 hCoV-19/USA/UT-UPHL-201008983/2020|EPI_ISL_602948|    ····················  ····················  ·····················  ····················  ····················  ····················
N    6627 hCoV-19/USA/UT-UPHL-201011997/2020|EPI_ISL_602949|    ····················  ····················  ·····················  ····················  ····················  ····················
N    6628 hCoV-19/USA/UT-UPHL-201005271/2020|EPI_ISL_602950|    ····················  ····················  ·····················  ····················  ····················  ····················
N    6629 hCoV-19/USA/UT-UPHL-201018562/2020|EPI_ISL_602951|    ····················  ····················  ·····················  ····················  ····················  ····················
N    6630 hCoV-19/USA/UT-UPHL-201022031/2020|EPI_ISL_602952|    ····················  ····················  ·····················  ····················  ····················  ····················
N    6631 hCoV-19/USA/UT-UPHL-201009564/2020|EPI_ISL_602955|    ····················  ····················  ·····················  ····················  ····················  ····················
N    6633 hCoV-19/USA/MN-MDH-1822/2020|EPI_ISL_602957|2020-0    ····················  ····················  ·····················  ····················  ····················  ····················
N    6634 hCoV-19/USA/MN-MDH-1823/2020|EPI_ISL_602958|2020-0    ····················  ····················  ·····················  ····················  ····················  ····················
N    6635 hCoV-19/USA/MN-MDH-1824/2020|EPI_ISL_602959|2020-0    ····················  ····················  ·····················  ····················  ····················  ····················
N    6636 hCoV-19/USA/MN-MDH-1825/2020|EPI_ISL_602960|2020-0    ····················  ·············A······  ·····················  ····················  ····················  ····················
N    6637 hCoV-19/USA/MN-MDH-1826/2020|EPI_ISL_602961|2020-0    ····················  ····················  ·····················  ····················  ····················  ····················
N    6638 hCoV-19/USA/MN-MDH-1827/2020|EPI_ISL_602962|2020-0    ····················  ····················  ·····················  ····················  ····················  ····················
N    6639 hCoV-19/USA/MN-MDH-1828/2020|EPI_ISL_602963|2020-0    ····················  ····················  ·····················  ····················  ····················  ····················
N    6640 hCoV-19/USA/MN-MDH-1829/2020|EPI_ISL_602964|2020-0    ····················  ····················  ·····················  ····················  ····················  ····················
N    6641 hCoV-19/USA/MN-MDH-1830/2020|EPI_ISL_602965|2020-0    ····················  ····················  ·····················  ····················  ····················  ····················
N    6642 hCoV-19/USA/MN-MDH-1831/2020|EPI_ISL_602966|2020-0    ····················  ····················  ·····················  ····················  ····················  ····················
N    6643 hCoV-19/USA/MN-MDH-1832/2020|EPI_ISL_602967|2020-0    ····················  ····················  ·····················  ····················  ····················  ····················
N    6644 hCoV-19/USA/MN-MDH-1833/2020|EPI_ISL_602968|2020-0    ····················  ····················  ·····················  ····················  ····················  ····················
N    6645 hCoV-19/USA/MN-MDH-1834/2020|EPI_ISL_602969|2020-0    ····················  ····················  ·····················  ····················  ····················  ····················
N    6646 hCoV-19/USA/MN-MDH-1835/2020|EPI_ISL_602970|2020-0    ····················  ····················  ·····················  ····················  ··N·················  ····················
N    6647 hCoV-19/USA/MN-MDH-1836/2020|EPI_ISL_602971|2020-0    ····················  ·············A······  ·····················  ····················  ····················  ····················
N    6648 hCoV-19/USA/MN-MDH-1837/2020|EPI_ISL_602972|2020-0    ····················  ····················  ·····················  ····················  ····················  ····················
N    6649 hCoV-19/USA/MN-MDH-1838/2020|EPI_ISL_602973|2020-0    ····················  ····················  ·····················  ····················  ····················  ····················
N    6650 hCoV-19/USA/MN-MDH-1839/2020|EPI_ISL_602974|2020-0    ····················  ····················  ·····················  ····················  ····················  ····················
N    6651 hCoV-19/USA/MN-MDH-1840/2020|EPI_ISL_602975|2020-0    ····················  ····················  ·····················  ····················  ····················  ····················
N    6652 hCoV-19/USA/MN-MDH-1841/2020|EPI_ISL_602976|2020-0    ····················  ·············A······  ·····················  ····················  ····················  ····················
N    6653 hCoV-19/USA/MN-MDH-1842/2020|EPI_ISL_602977|2020-0    ····················  ····················  ·····················  ····················  ····················  ····················
N    6654 hCoV-19/USA/MN-MDH-1855/2020|EPI_ISL_602990|2020-1    ····················  ·A··················  ·····················  ····················  ····················  ····················
N    6655 hCoV-19/USA/MN-MDH-1856/2020|EPI_ISL_602991|2020-1    ····················  ·A··················  ·····················  ····················  ····················  ····················
N    6656 hCoV-19/USA/MN-MDH-1860/2020|EPI_ISL_602995|2020-1    ····················  ·A··················  ·····················  ····················  ····················  ····················
N    6657 hCoV-19/Palestine/39/2020|EPI_ISL_596521|2020-07-2    ····················  ····················  ·····················  ····················  ····················  ····················
N    6658 hCoV-19/Palestine/40/2020|EPI_ISL_596523|2020-08-1    ····················  ····················  ·····················  ····················  ····················  ····················
N    6659 hCoV-19/Palestine/42/2020|EPI_ISL_596525|2020-08-1    ····················  ····················  ·····················  ····················  ····················  ····················
N    6660 hCoV-19/Palestine/43/2020|EPI_ISL_596526|2020-08-1    ····················  ····················  ·····················  ····················  ····················  ····················
N    6661 hCoV-19/Palestine/44/2020|EPI_ISL_596527|2020-08-1    ····················  ····················  ·····················  ····················  ····················  ····················
N    6662 hCoV-19/Palestine/45/2020|EPI_ISL_596528|2020-08-1    ····················  ····················  ·····················  ····················  ····················  ····················
N    6663 hCoV-19/Palestine/46/2020|EPI_ISL_596529|2020-08-1    ····················  ····················  ·····················  ····················  ····················  ····················
N    6664 hCoV-19/Palestine/47/2020|EPI_ISL_596530|2020-08-1    ····················  ····················  ·····················  ····················  ····················  ····················
N    6665 hCoV-19/Palestine/48/2020|EPI_ISL_596531|2020-08-1    ····················  ····················  ·····················  ····················  ····················  ····················
N    6666 hCoV-19/Palestine/49/2020|EPI_ISL_596532|2020-08-1    ····················  ····················  ·····················  ····················  ····················  ····················
N    6667 hCoV-19/Palestine/4/2020|EPI_ISL_596533|2020-07-17    ····················  ····················  ·····················  ····················  ····················  ····················
N    6668 hCoV-19/Palestine/50/2020|EPI_ISL_596534|2020-08-1    ····················  ····················  ·····················  ····················  ····················  ····················
N    6669 hCoV-19/Palestine/51/2020|EPI_ISL_596535|2020-08-1    ····················  ····················  ·····················  ····················  ····················  ····················
N    6670 hCoV-19/Palestine/52/2020|EPI_ISL_596536|2020-08-1    ····················  ····················  ·····················  ····················  ····················  ····················
N    6671 hCoV-19/Palestine/54/2020|EPI_ISL_596537|2020-08-1    ····················  ····················  ·····················  ····················  ····················  ····················
N    6672 hCoV-19/Palestine/55/2020|EPI_ISL_596538|2020-08-1    ····················  ····················  ·····················  ····················  ····················  ····················
N    6673 hCoV-19/Palestine/60/2020|EPI_ISL_596539|2020-03      ····················  ····················  ·····················  ····················  ····················  ····················
N    6674 hCoV-19/Palestine/61/2020|EPI_ISL_596540|2020-03      ····················  ····················  ·····················  ····················  ····················  ····················
N    6675 hCoV-19/Palestine/62/2020|EPI_ISL_596541|2020-03      ····················  ····················  ·····················  ····················  ····················  ····················
```

N 6676 hCoV-19/Palestine/64/2020|EPI_ISL_596542|2020-06-2 ···················· ···················· ····················· ···················· ···················· ····················
N 6677 hCoV-19/USA/MI-UM-10036276912/2020|EPI_ISL_596588| ···················· ·············A······ ····················· ···················· ···················· ····················
N 6678 hCoV-19/USA/MI-UM-10036305857/2020|EPI_ISL_596589| ···················· ···················· ····················· ···················· ···················· ····················
N 6679 hCoV-19/USA/MI-UM-10036282773/2020|EPI_ISL_596593| ···················· ···················· ····················· ···················· ···················· ····················
N 6680 hCoV-19/USA/MI-UM-10036319590/2020|EPI_ISL_596594| ···················· ···················· ····················· ···················· ···················· ····················
N 6681 hCoV-19/USA/MI-UM-10036296398/2020|EPI_ISL_596596| ···················· ···················· ····················· ···················· ···················· ····················
N 6682 hCoV-19/USA/MI-UM-10036284170/2020|EPI_ISL_596597| ···················· ···················· ····················· ···················· ···················· ····················
N 6683 hCoV-19/USA/MI-UM-10036306472/2020|EPI_ISL_596598| ···················· ···················· ····················· ···················· ···················· ····················
N 6684 hCoV-19/USA/MI-UM-10036312721/2020|EPI_ISL_596599| ···················· ···················· ····················· ···················· ···················· ····················
N 6685 hCoV-19/England/ALDP-A5090C/2020|EPI_ISL_597800|20 ···················· ···················· ····················· ···················· ···················· ····················
N 6686 hCoV-19/England/ALDP-A5075D/2020|EPI_ISL_597801|20 ···················· ···················· ····················· ···················· ···················· ····················
N 6687 hCoV-19/England/ALDP-A50FA0/2020|EPI_ISL_597802|20 ···················· ···················· ····················· ···················· ···················· ····················
N 6688 hCoV-19/England/ALDP-A50A54/2020|EPI_ISL_597803|20 ···················· ···················· ····················· ···················· ···················· ····················
N 6689 hCoV-19/England/ALDP-A50B8E/2020|EPI_ISL_597804|20 ···················· ···················· ····················· ···················· ···················· ····················
N 6690 hCoV-19/England/ALDP-A50948/2020|EPI_ISL_597805|20 ···················· ···················· ····················· ···················· ···················· ····················
N 6691 hCoV-19/England/ALDP-A4F9F1/2020|EPI_ISL_597806|20 ···················· ···················· ····················· ···················· ···················· ····················
N 6692 hCoV-19/England/ALDP-A50B33/2020|EPI_ISL_597807|20 ···················· ···················· ····················· ···················· ···················· ····················
N 6693 hCoV-19/England/ALDP-A50070/2020|EPI_ISL_597808|20 ···················· ···················· ····················· ···················· ···················· ····················
N 6694 hCoV-19/England/ALDP-A51033/2020|EPI_ISL_597809|20 ···················· ···················· ····················· ···················· ···················· ····················
N 6695 hCoV-19/England/ALDP-A5108E/2020|EPI_ISL_597820|20 ···················· ·A·················· ····················· ···················· ···················· ····················
N 6696 hCoV-19/England/ALDP-A50B7F/2020|EPI_ISL_597822|20 ···················· ···················· ····················· ···················· ···················· ····················
N 6697 hCoV-19/England/ALDP-A5082D/2020|EPI_ISL_597823|20 ···················· ···················· ····················· ···················· ···················· ····················
N 6698 hCoV-19/England/ALDP-A4FE01/2020|EPI_ISL_597824|20 ···················· ···················· ····················· ···················· ···················· ····················
N 6699 hCoV-19/England/ALDP-A50C7C/2020|EPI_ISL_597825|20 ···················· ···················· ····················· ···················· ···················· ····················
N 6700 hCoV-19/England/ALDP-A5041A/2020|EPI_ISL_597826|20 ···················· ···················· ····················· ···················· ···················· ····················
N 6701 hCoV-19/England/ALDP-A510AC/2020|EPI_ISL_597852|20 ···················· ···················· ····················· ···················· ···················· ····················
N 6702 hCoV-19/England/ALDP-A5032C/2020|EPI_ISL_597853|20 ···················· ···················· ····················· ···················· ···················· ····················
N 6703 hCoV-19/England/ALDP-A5092A/2020|EPI_ISL_597854|20 ···················· ···················· ····················· ···················· ···················· ····················
N 6704 hCoV-19/England/ALDP-A5015F/2020|EPI_ISL_597855|20 ···················· ···················· ····················· ···················· ···················· ····················
N 6705 hCoV-19/England/ALDP-A50A09/2020|EPI_ISL_597856|20 ···················· ···················· ····················· ···················· ···················· ····················
N 6706 hCoV-19/England/ALDP-A4F988/2020|EPI_ISL_597857|20 ···················· ···················· ····················· ···················· ···················· ····················
N 6707 hCoV-19/England/ALDP-A5019B/2020|EPI_ISL_597858|20 ···················· ···················· ····················· ···················· ···················· ····················
N 6708 hCoV-19/England/ALDP-A4FDAA/2020|EPI_ISL_597859|20 ···················· ···················· ····················· ···················· ···················· ····················
N 6709 hCoV-19/England/ALDP-A51042/2020|EPI_ISL_597860|20 ···················· ···················· ····················· ···················· ···················· ····················
N 6710 hCoV-19/England/ALDP-A50BD9/2020|EPI_ISL_597861|20 ···················· ···················· ····················· ···················· ···················· ····················
N 6711 hCoV-19/England/ALDP-A501F5/2020|EPI_ISL_597862|20 ···················· ···················· ····················· ···················· ···················· ····················
N 6712 hCoV-19/England/ALDP-A50EC1/2020|EPI_ISL_597864|20 ···················· ···················· ····················· ···················· ···················· ····················
N 6713 hCoV-19/England/ALDP-A5074E/2020|EPI_ISL_597865|20 ···················· ···················· ····················· ···················· ···················· ····················
N 6714 hCoV-19/England/ALDP-A4FA76/2020|EPI_ISL_597866|20 ···················· ···················· ····················· ···················· ···················· ····················
N 6715 hCoV-19/England/ALDP-A50043/2020|EPI_ISL_597867|20 ···················· ···················· ····················· ···················· ···················· ····················
N 6716 hCoV-19/England/ALDP-A50140/2020|EPI_ISL_597868|20 ···················· ···················· ····················· ···················· ···················· ····················
N 6717 hCoV-19/England/ALDP-A50966/2020|EPI_ISL_597869|20 ···················· ···················· ····················· ···················· ···················· ····················
N 6718 hCoV-19/England/ALDP-A50E76/2020|EPI_ISL_597870|20 ···················· ···················· ····················· ···················· ···················· ····················
N 6719 hCoV-19/England/ALDP-A502E3/2020|EPI_ISL_597871|20 ···················· ···················· ····················· ···················· ···················· ····················
N 6720 hCoV-19/England/ALDP-A50E1C/2020|EPI_ISL_597872|20 ···················· ···················· ····················· ···················· ···················· ····················
N 6721 hCoV-19/England/ALDP-A50B9D/2020|EPI_ISL_597873|20 ···················· ···················· ····················· ···················· ···················· ····················
N 6722 hCoV-19/England/QEUH-A008D1/2020|EPI_ISL_600200|20 ···················· ···················· ····················· ···················· ···················· ····················
N 6723 hCoV-19/England/QEUH-9FF997/2020|EPI_ISL_600201|20 ···················· ···················· ····················· ···················· ···················· ····················
N 6724 hCoV-19/Scotland/QEUH-A007E3/2020|EPI_ISL_600203|2 ···················· ···················· ····················· ···················· ···················· ····················
N 6725 hCoV-19/England/QEUH-9FFAB2/2020|EPI_ISL_600204|20 ···················· ···················· ····················· ···················· ···················· ····················
N 6726 hCoV-19/Scotland/QEUH-A0066E/2020|EPI_ISL_600205|2 ···················· ···················· ····················· ···················· ···················· ····················
N 6727 hCoV-19/Scotland/QEUH-9FF78E/2020|EPI_ISL_600207|2 ···················· ···················· ····················· ···················· ···················· ····················
N 6728 hCoV-19/England/QEUH-A00349/2020|EPI_ISL_600208|20 ···················· ···················· ····················· ···················· ···················· ····················
N 6729 hCoV-19/England/QEUH-A008E0/2020|EPI_ISL_600209|20 ···················· ·A·················· ····················· ···················· ···················· ····················
N 6730 hCoV-19/England/QEUH-9FF3C6/2020|EPI_ISL_600210|20 ···················· ·A·················· ····················· ···················· ···················· ····················
N 6731 hCoV-19/England/QEUH-9FFA76/2020|EPI_ISL_600212|20 ···················· ···················· ····················· ···················· ······T············· ····················
N 6732 hCoV-19/Scotland/QEUH-9FFF1D/2020|EPI_ISL_600213|2 ···················· ···················· ····················· ···················· ···················· ····················
N 6733 hCoV-19/Scotland/QEUH-9FFE89/2020|EPI_ISL_600214|2 ···················· ···················· ····················· ···················· ···················· ····················
N 6734 hCoV-19/England/QEUH-9FF41E/2020|EPI_ISL_600215|20 ···················· ···················· ····················· ···················· ······T············· ····················
N 6735 hCoV-19/England/QEUH-A0077A/2020|EPI_ISL_600216|20 ···················· ···················· ····················· ···················· ···················· ····················
N 6736 hCoV-19/England/QEUH-A0032B/2020|EPI_ISL_600218|20 ···················· ···················· ····················· ···················· ····T··············· ····················
N 6737 hCoV-19/England/QEUH-9FF232/2020|EPI_ISL_600219|20 ···················· ···················· ····················· ···················· ···················· ····················
N 6738 hCoV-19/England/QEUH-A00929/2020|EPI_ISL_600250|20 ···················· ···················· ····················· ···················· ···················· ····················
N 6739 hCoV-19/England/QEUH-9FF6FA/2020|EPI_ISL_600251|20 ···················· ···················· ····················· ···················· ···················· ····················
N 6740 hCoV-19/England/QEUH-A00956/2020|EPI_ISL_600257|20 ···················· ···················· ····················· ···················· ······T············· ····················
N 6741 hCoV-19/Wales/QEUH-A0017C/2020|EPI_ISL_600258|2020 ···················· ···················· ····················· ···················· ······T············· ····················
N 6742 hCoV-19/England/QEUH-9FF399/2020|EPI_ISL_600259|20 ···················· ···················· ····················· ···················· ···················· ····················
N 6743 hCoV-19/England/QEUH-9FFB46/2020|EPI_ISL_600261|20 ···················· ···················· ····················· ···················· ···················· ····················
N 6744 hCoV-19/England/QEUH-A00947/2020|EPI_ISL_600262|20 ···················· ···················· ····················· ···················· ···················· ····················
N 6745 hCoV-19/England/QEUH-9FFBEC/2020|EPI_ISL_600264|20 ···················· ···················· ····················· ···················· ···················· ····················
N 6746 hCoV-19/England/QEUH-9FF6DC/2020|EPI_ISL_600265|20 ···················· ···················· ····················· ···················· ···················· ····················
N 6747 hCoV-19/England/QEUH-A00710/2020|EPI_ISL_600266|20 ···················· ···················· ····················· ···················· ···················· ····················
N 6748 hCoV-19/England/QEUH-A0083B/2020|EPI_ISL_600267|20 ···················· ·A·················· ····················· ···················· ···················· ···A················
N 6749 hCoV-19/Scotland/QEUH-9FF9D3/2020|EPI_ISL_600268|2 ···················· ···················· ····················· ···················· ···················· ····················
N 6750 hCoV-19/Scotland/QEUH-9FFA67/2020|EPI_ISL_600269|2 ···················· ···················· ····················· ···················· ···················· ····················
N 6751 hCoV-19/England/QEUH-A00419/2020|EPI_ISL_600270|20 ···················· ···················· ····················· ···················· ···················· ····················
N 6752 hCoV-19/England/QEUH-9FF988/2020|EPI_ISL_600271|20 ···················· ···················· ····················· ···················· ···················· ····················
N 6753 hCoV-19/England/QEUH-A0026A/2020|EPI_ISL_600272|20 ···················· ·A·················· ····················· ···················· ···················· ···A················
N 6754 hCoV-19/England/QEUH-A00051/2020|EPI_ISL_600274|20 ···················· ···················· ····················· ···················· ···················· ····················
N 6755 hCoV-19/England/QEUH-A002A6/2020|EPI_ISL_600275|20 ···················· ···················· ····················· ···················· ······T············· ····················

N 6756 hCoV-19/England/QEUH-9FF37B/2020|EPI_ISL_600276|20 ···················· ···················· ····················· ···················· ···················· ····················
N 6757 hCoV-19/England/QEUH-A002C4/2020|EPI_ISL_600277|20 ···················· ···················· ····················· ···················· ···················· ····················
N 6758 hCoV-19/England/QEUH-9FF1EA/2020|EPI_ISL_600278|20 ···················· ···················· ····················· ···················· ···················· ····················
N 6759 hCoV-19/England/QEUH-9FFB28/2020|EPI_ISL_600279|20 ···················· ···················· ····················· ···················· ···················· ····················
N 6760 hCoV-19/England/MILK-9D4F3C/2020|EPI_ISL_601526|20 ···················· ···················· ····················· ···················· ···················· ····················
N 6761 hCoV-19/Wales/MILK-9D4CEA/2020|EPI_ISL_601527|2020 ···················· ···················· ····················· ···················· ···················· ····················
N 6762 hCoV-19/England/MILK-9D3EA9/2020|EPI_ISL_601529|20 ···················· ·A·················· ····················· ···················· ···················· ···A················
N 6763 hCoV-19/England/CAMC-9D1F4E/2020|EPI_ISL_601530|20 ···················· ···················· ····················· ···················· ···················· ····················
N 6764 hCoV-19/England/MILK-9D4F2D/2020|EPI_ISL_601531|20 ···················· ···················· ····················· ···················· ···················· ····················
N 6765 hCoV-19/England/MILK-9D4DF6/2020|EPI_ISL_601532|20 ···················· C··················· ····················· ···················· ···················· ····················
N 6766 hCoV-19/England/MILK-9D4D7E/2020|EPI_ISL_601533|20 ···················· ···················· ····················· ···················· ···················· ····················
N 6767 hCoV-19/England/MILK-9D3B48/2020|EPI_ISL_601534|20 ···················· ···················· ····················· ···················· ···················· ····················
N 6768 hCoV-19/England/MILK-9D4A59/2020|EPI_ISL_601535|20 ···················· ···················· ····················· ···················· ···················· ····················
N 6769 hCoV-19/England/MILK-9D3DBB/2020|EPI_ISL_601537|20 ···················· ···················· ····················· ···················· ···················· ····················
N 6770 hCoV-19/Wales/MILK-9D45B2/2020|EPI_ISL_601538|2020 ···················· ···················· ····················· ···················· ···················· ····················
N 6771 hCoV-19/England/MILK-9D4FD2/2020|EPI_ISL_601539|20 ···················· ···················· ····················· ···················· ···················· ····················
N 6772 hCoV-19/England/MILK-9D4558/2020|EPI_ISL_601540|20 ···················· ···················· ····················· ···················· ···················· ····················
N 6773 hCoV-19/England/CAMC-9D2217/2020|EPI_ISL_601541|20 ···················· ···················· ····················· ···················· ···················· ····················
N 6774 hCoV-19/Wales/MILK-9D4D8D/2020|EPI_ISL_601542|2020 ···················· ···················· ····················· ···················· ···················· ····················
N 6775 hCoV-19/England/MILK-9D4F1E/2020|EPI_ISL_601543|20 ···················· ···················· ····················· ···················· ···················· ····················
N 6776 hCoV-19/England/MILK-9D488C/2020|EPI_ISL_601545|20 ···················· ···················· ····················· ···················· ···················· ····················
N 6777 hCoV-19/England/MILK-9D489B/2020|EPI_ISL_601546|20 ···················· ···················· ····················· ···················· ···················· ····················
N 6778 hCoV-19/England/CAMC-9D20E0/2020|EPI_ISL_601580|20 ···················· ···················· ····················· ···················· ···················· ····················
N 6779 hCoV-19/England/MILK-9D4655/2020|EPI_ISL_601581|20 ···················· ···················· ····················· ···················· ···················· ····················
N 6780 hCoV-19/England/MILK-9D4761/2020|EPI_ISL_601582|20 ···················· ·A·················· ····················· ···················· ···················· ····················
N 6781 hCoV-19/England/MILK-9D485F/2020|EPI_ISL_601583|20 ···················· ···················· ····················· ···················· ···················· ····················
N 6782 hCoV-19/Wales/CAMC-9D2350/2020|EPI_ISL_601588|2020 ···················· ···················· ····················· ···················· ···················· ····················
N 6783 hCoV-19/England/CAMC-9D1CB0/2020|EPI_ISL_601589|20 ···················· ···················· ····················· ···················· ···················· ····················
N 6784 hCoV-19/England/CAMC-9D21ED/2020|EPI_ISL_601591|20 ···················· ···················· ····················· ···················· ····T··············· ····················
N 6785 hCoV-19/England/MILK-9D4549/2020|EPI_ISL_601592|20 ···················· ···················· ····················· ···················· ···················· ····················
N 6786 hCoV-19/Wales/MILK-9D3FC4/2020|EPI_ISL_601593|2020 ···················· ···················· ····················· ···················· ···················· ····················
N 6787 hCoV-19/England/CAMC-9D2156/2020|EPI_ISL_601594|20 ···················· ···················· ····················· ···················· ···················· ····················
N 6788 hCoV-19/England/MILK-9D4E3F/2020|EPI_ISL_601596|20 ···················· ···················· ····················· ···················· ······T············· ····················
N 6789 hCoV-19/England/MILK-9D47F8/2020|EPI_ISL_601598|20 ···················· ···················· ····················· ···················· ···················· ····················
N 6790 hCoV-19/England/MILK-9D4770/2020|EPI_ISL_601599|20 ···················· ···················· ····················· ···················· ······T············· ····················
N 6791 hCoV-19/South Africa/KRISP-K003466/2020|EPI_ISL_60 ···················· ···················· ····················· ···················· ···················· ····················
N 6792 hCoV-19/South Africa/KRISP-K003467/2020|EPI_ISL_60 ···················· ·A·················· ····················· ···················· ···················· ····················
N 6793 hCoV-19/South Africa/KRISP-K003469/2020|EPI_ISL_60 ···················· ···················· ····················· ···················· ···················· ····················
N 6794 hCoV-19/South Africa/KRISP-K003470/2020|EPI_ISL_60 ···················· ·A·················· ····················· ···················· ···················· ····················
N 6795 hCoV-19/South Africa/KRISP-K003471/2020|EPI_ISL_60 ···················· ·A·················· ····················· ···················· ···················· ····················
N 6796 hCoV-19/South Africa/KRISP-K003473/2020|EPI_ISL_60 ···················· ···················· ····················· ···················· ···················· ····················
N 6797 hCoV-19/South Africa/KRISP-K003474/2020|EPI_ISL_60 ···················· ···················· ····················· ···················· ···················· ····················
N 6798 hCoV-19/South Africa/KRISP-K003476/2020|EPI_ISL_60 ···················· ···················· ····················· ···················· ···················· ····················
N 6799 hCoV-19/South Africa/KRISP-K003477/2020|EPI_ISL_60 ···················· ···················· ····················· ···················· ···················· ····················
N 6800 hCoV-19/South Africa/KRISP-K003478/2020|EPI_ISL_60 ···················· ···················· ····················· ···················· ···················· ····················
N 6801 hCoV-19/South Africa/KRISP-K003479/2020|EPI_ISL_60 ···················· ···················· ····················· ···················· ···················· ····················
N 6802 hCoV-19/South Africa/KRISP-K003480/2020|EPI_ISL_60 ···················· ···················· ····················· ···················· ···················· ····················
N 6803 hCoV-19/South Africa/KRISP-K003541/2020|EPI_ISL_60 ···················· ···················· ····················· ···················· ···················· ····················
N 6804 hCoV-19/South Africa/KRISP-K003543/2020|EPI_ISL_60 ···················· ···················· ····················· ···················· ···················· ····················
N 6805 hCoV-19/South Africa/KRISP-K003544/2020|EPI_ISL_60 ···················· ···················· ····················· ···················· ···················· ····················
N 6806 hCoV-19/South Africa/KRISP-K003545/2020|EPI_ISL_60 ···················· ···················· ····················· ···················· ···················· ····················
N 6807 hCoV-19/South Africa/KRISP-K003546/2020|EPI_ISL_60 ···················· ···················· ····················· ···················· ···················· ····················
N 6808 hCoV-19/South Africa/KRISP-K003547/2020|EPI_ISL_60 ···················· ···················· ····················· ···················· ···················· ····················
N 6809 hCoV-19/South Africa/KRISP-K003550/2020|EPI_ISL_60 ···················· ···················· ····················· ···················· ···················· ····················
N 6810 hCoV-19/South Africa/KRISP-K003679/2020|EPI_ISL_60 ···················· ··········A········· ····················· ···················· ···················· ····················
N 6811 hCoV-19/South Africa/KRISP-K003683/2020|EPI_ISL_60 ···················· ···················· ····················· ···················· ···················· ····················
N 6812 hCoV-19/South Africa/KRISP-K003684/2020|EPI_ISL_60 ···················· ···················· ····················· ···················· ···················· ····················
N 6814 hCoV-19/South Africa/KRISP-K003688/2020|EPI_ISL_60 ···················· ···················· ····················· ···················· ···················· ····················
N 6815 hCoV-19/South Africa/KRISP-K003689/2020|EPI_ISL_60 ···················· ···················· ····················· ···················· ···················· ····················
N 6816 hCoV-19/South Africa/KRISP-K003690/2020|EPI_ISL_60 ···················· ···················· ····················· ···················· ···················· ····················
N 6817 hCoV-19/South Africa/KRISP-K003691/2020|EPI_ISL_60 ···················· ···················· ····················· ···················· ···················· ····················
N 6818 hCoV-19/South Africa/KRISP-K003694/2020|EPI_ISL_60 ···················· ·A·················· ····················· ···················· ···················· ····················
N 6819 hCoV-19/South Africa/KRISP-K003696/2020|EPI_ISL_60 ···················· ···················· ····················· ···················· ···················· ····················
N 6821 hCoV-19/South Africa/KRISP-K003698/2020|EPI_ISL_60 ···················· ···················· ····················· ···················· ···················· ····················
N 6822 hCoV-19/USA/CA-QDX-2720/2020|EPI_ISL_605010|2020-0 ···················· ···················· ····················· ···················· ···················· ····················
N 6823 hCoV-19/USA/CA-QDX-2721/2020|EPI_ISL_605011|2020-0 ···················· ···················· ····················· ···················· ···················· ····················
N 6824 hCoV-19/USA/CA-QDX-2724/2020|EPI_ISL_605012|2020-0 ···················· ···················· ····················· ···················· ···················· ····················
N 6825 hCoV-19/USA/CA-QDX-2728/2020|EPI_ISL_605013|2020-0 ···················· ···················· ····················· ···················· ···················· ····················
N 6826 hCoV-19/USA/CA-QDX-2733/2020|EPI_ISL_605014|2020-0 ···················· ···················· ····················· ···················· ···················· ····················
N 6827 hCoV-19/USA/CA-QDX-2737/2020|EPI_ISL_605015|2020-0 ···················· ···················· ····················· ···················· ···················· ····················
N 6828 hCoV-19/USA/CA-QDX-2739/2020|EPI_ISL_605016|2020-0 ···················· ···················· ····················· ···················· ···················· ····················
N 6829 hCoV-19/USA/CA-QDX-2741/2020|EPI_ISL_605017|2020-0 ···················· ···················· ····················· ···················· ···················· ····················
N 6830 hCoV-19/USA/CA-QDX-2742/2020|EPI_ISL_605018|2020-0 ···················· ···················· ····················· ···················· ···················· ····················
N 6831 hCoV-19/USA/CA-QDX-2743/2020|EPI_ISL_605019|2020-0 ···················· ···················· ····················· ···················· ···················· ····················
N 6832 hCoV-19/USA/CA-QDX-2744/2020|EPI_ISL_605020|2020-0 ···················· ···················· ····················· ···················· ···················· ····················
N 6833 hCoV-19/USA/CA-QDX-2745/2020|EPI_ISL_605021|2020-0 ···················· ···················· ····················· ···················· ···················· ····················
N 6834 hCoV-19/USA/CA-QDX-2746/2020|EPI_ISL_605022|2020-0 ···················· ···················· ····················· ···················· ···················· ····················
N 6835 hCoV-19/USA/CT-QDX-2695/2020|EPI_ISL_605023|2020-0 ···················· ···················· ····················· ···················· ···················· ····················
N 6836 hCoV-19/USA/MA-QDX-2690/2020|EPI_ISL_605024|2020-0 ···················· ···················· ····················· ···················· ···················· ····················
N 6837 hCoV-19/USA/MA-QDX-2708/2020|EPI_ISL_605025|2020-0 ···················· ···················· ····················· ···················· ···················· ····················

| | | | | | | | | |
|---|---|---|---|---|---|---|---|---|
| N | 6838 hCoV-19/USA/MN-QDX-2704/2020\|EPI_ISL_605026\|2020-0 | ···················· | ···················· | ···················· | ···················· | ···················· | ···················· | |
| N | 6839 hCoV-19/USA/MN-QDX-2715/2020\|EPI_ISL_605027\|2020-0 | ···················· | ···················· | ···················· | ···················· | ···················· | ···················· | |
| N | 6840 hCoV-19/USA/MN-QDX-2716/2020\|EPI_ISL_605028\|2020-0 | ···················· | ···················· | ···················· | ···················· | ···················· | ···················· | |
| N | 6841 hCoV-19/USA/MN-QDX-2717/2020\|EPI_ISL_605029\|2020-0 | ···················· | ···················· | ···················· | ···················· | ···················· | ···················· | |
| N | 6842 hCoV-19/USA/MN-QDX-2719/2020\|EPI_ISL_605030\|2020-0 | ···················· | ···················· | ···················· | ···················· | ···················· | ···················· | |
| N | 6843 hCoV-19/USA/MN-QDX-2725/2020\|EPI_ISL_605031\|2020-0 | ···················· | ···················· | ···················· | ···················· | ···················· | ···················· | |
| N | 6844 hCoV-19/USA/MN-QDX-2726/2020\|EPI_ISL_605032\|2020-0 | ···················· | ···················· | ···················· | ···················· | ···················· | ···················· | |
| N | 6845 hCoV-19/USA/MN-QDX-2727/2020\|EPI_ISL_605033\|2020-0 | ···················· | ···················· | ···················· | ···················· | ···················· | ···················· | |
| N | 6846 hCoV-19/USA/MN-QDX-2730/2020\|EPI_ISL_605034\|2020-0 | ···················· | ···················· | ···················· | ···················· | ···················· | ···················· | |
| N | 6847 hCoV-19/USA/MN-QDX-2736/2020\|EPI_ISL_605035\|2020-0 | ···················· | ···················· | ···················· | ···················· | ···················· | ···················· | |
| N | 6848 hCoV-19/USA/MN-QDX-2738/2020\|EPI_ISL_605036\|2020-0 | ···················· | ···················· | ···················· | ···················· | ···················· | ···················· | |
| N | 6849 hCoV-19/USA/NV-QDX-2723/2020\|EPI_ISL_605037\|2020-0 | ···················· | ···················· | ···················· | ···················· | ···················· | ···················· | |
| N | 6850 hCoV-19/USA/NY-QDX-2682/2020\|EPI_ISL_605038\|2020-0 | ···················· | ···················· | ···················· | ···················· | ···················· | ···················· | |
| N | 6851 hCoV-19/USA/NY-QDX-2689/2020\|EPI_ISL_605039\|2020-0 | ···················· | ···················· | ···················· | ···················· | ···················· | ···················· | |
| N | 6852 hCoV-19/USA/NY-QDX-2693/2020\|EPI_ISL_605040\|2020-0 | ···················· | ···················· | ···················· | ···················· | ···················· | ···················· | |
| N | 6853 hCoV-19/USA/NY-QDX-2696/2020\|EPI_ISL_605041\|2020-0 | ···················· | ···················· | ···················· | ···················· | ···················· | ···················· | |
| N | 6854 hCoV-19/USA/NY-QDX-2698/2020\|EPI_ISL_605042\|2020-0 | ···················· | ···················· | ···················· | ···················· | ···················· | ···················· | |
| N | 6855 hCoV-19/USA/NY-QDX-2701/2020\|EPI_ISL_605043\|2020-0 | ···················· | ···················· | ···················· | ···················· | ···················· | ···················· | |
| N | 6856 hCoV-19/USA/NY-QDX-2702/2020\|EPI_ISL_605044\|2020-0 | ···················· | ···················· | ···················· | ···················· | ···················· | ···················· | |
| N | 6857 hCoV-19/USA/NY-QDX-2706/2020\|EPI_ISL_605045\|2020-0 | ···················· | ···················· | ···················· | ···················· | ···················· | ···················· | |
| N | 6858 hCoV-19/USA/NY-QDX-2707/2020\|EPI_ISL_605046\|2020-0 | ···················· | ···················· | ···················· | ···················· | ···················· | ···················· | |
| N | 6859 hCoV-19/USA/NY-QDX-2711/2020\|EPI_ISL_605047\|2020-0 | ···················· | ···················· | ···················· | ···················· | ···················· | ···················· | |
| N | 6860 hCoV-19/USA/NY-QDX-2712/2020\|EPI_ISL_605048\|2020-0 | ···················· | ···················· | ···················· | ···················· | ···················· | ···················· | |
| N | 6861 hCoV-19/USA/NY-QDX-2714/2020\|EPI_ISL_605049\|2020-0 | ···················· | ···················· | ···················· | ···················· | ···················· | ···················· | |
| N | 6862 hCoV-19/USA/NY-QDX-2722/2020\|EPI_ISL_605050\|2020-0 | ···················· | ···················· | ···················· | ···················· | ···················· | ···················· | |
| N | 6863 hCoV-19/USA/NY-QDX-2729/2020\|EPI_ISL_605051\|2020-0 | ···················· | ···················· | ···················· | ···················· | ···················· | ···················· | |
| N | 6864 hCoV-19/USA/NY-QDX-2731/2020\|EPI_ISL_605052\|2020-0 | ···················· | ···················· | ···················· | ···················· | ···················· | ···················· | |
| N | 6865 hCoV-19/USA/NY-QDX-2732/2020\|EPI_ISL_605053\|2020-0 | ···················· | ···················· | ···················· | ···················· | ···················· | ···················· | |
| N | 6866 hCoV-19/USA/NY-QDX-2734/2020\|EPI_ISL_605054\|2020-0 | ···················· | ···················· | ···················· | ···················· | ···················· | ···················· | |
| N | 6867 hCoV-19/USA/NY-QDX-2735/2020\|EPI_ISL_605055\|2020-0 | ···················· | ···················· | ···················· | ···················· | ···················· | ···················· | |
| N | 6868 hCoV-19/USA/NY-QDX-2740/2020\|EPI_ISL_605056\|2020-0 | ···················· | ···················· | ···················· | ···················· | ···················· | ···················· | |
| N | 6869 hCoV-19/USA/TN-QDX-2686/2020\|EPI_ISL_605057\|2020-0 | ···················· | ···················· | ···················· | ···················· | ···················· | ···················· | |
| N | 6870 hCoV-19/Ireland/D-NVRL-20G44567/2020\|EPI_ISL_60505 | ···················· | ···················· | ···················· | ···················· | ···················· | ···················· | |
| N | 6871 hCoV-19/Ireland/D-NVRL-20G35361/2020\|EPI_ISL_60506 | ···················· | ···················· | ···················· | ···················· | ···················· | ···················· | |
| N | 6872 hCoV-19/Ireland/D-NVRL-20G44451/2020\|EPI_ISL_60506 | ···················· | ···················· | ···················· | ···················· | ···················· | ···················· | |
| N | 6873 hCoV-19/Ireland/D-NVRL-20G44520/2020\|EPI_ISL_60506 | ···················· | ···················· | ···················· | ···················· | ···················· | ···················· | |
| N | 6874 hCoV-19/Ireland/D-NVRL-20G44565/2020\|EPI_ISL_60506 | ···················· | ···················· | ···················· | ···················· | ···················· | ···················· | |
| N | 6875 hCoV-19/Ireland/D-NVRL-20G31911/2020\|EPI_ISL_60506 | ···················· | ···················· | ···················· | ···················· | ···················· | ···················· | |
| N | 6876 hCoV-19/Ireland/D-NVRL-20G44446/2020\|EPI_ISL_60506 | ···················· | ···················· | ···················· | ···················· | ···················· | ···················· | |
| N | 6877 hCoV-19/Ireland/D-NVRL-20G44517/2020\|EPI_ISL_60506 | ···················· | ···················· | ···················· | ···················· | ···················· | ···················· | |
| N | 6878 hCoV-19/Ireland/D-NVRL-20G44563/2020\|EPI_ISL_60506 | ···················· | ···················A | ···················· | ···················· | ···················· | ···················· | 1 |
| N | 6879 hCoV-19/Ireland/D-NVRL-20G35359/2020\|EPI_ISL_60506 | ···················· | ···················· | ···················· | ···················· | ···················· | ···················· | |
| N | 6880 hCoV-19/Ireland/D-NVRL-20G44438/2020\|EPI_ISL_60507 | ···················· | ···················· | ···················· | ···················· | ···················· | ···················· | |
| N | 6881 hCoV-19/Ireland/D-NVRL-20G44549/2020\|EPI_ISL_60507 | ···················· | ···················· | ···················· | ···················· | ···················· | ···················· | |
| N | 6882 hCoV-19/Ireland/MO-NVRL-20G31886/2020\|EPI_ISL_6050 | ···················· | ···················· | ···················· | ···················· | ···················· | ···················· | |
| N | 6883 hCoV-19/Ireland/WW-NVRL-20G35325/2020\|EPI_ISL_6050 | ···················· | ···················· | ···················· | ···················· | ···················· | ···················· | |
| N | 6884 hCoV-19/Ireland/D-NVRL-20G44426/2020\|EPI_ISL_60507 | ···················· | ···················· | ···················· | ···················· | ···················· | ···················· | |
| N | 6885 hCoV-19/Ireland/WW-NVRL-20G41355/2020\|EPI_ISL_6050 | ···················· | ···················· | ···················· | ···················· | ···················· | ···················· | |
| N | 6886 hCoV-19/Ireland/D-NVRL-20G44546/2020\|EPI_ISL_60507 | ···················· | ···················· | ···················· | ···················· | ···················· | ···················· | |
| N | 6887 hCoV-19/Ireland/OY-NVRL-20G31858/2020\|EPI_ISL_6050 | ···················· | ···················· | ···················· | ···················· | ···················· | ···················· | |
| N | 6888 hCoV-19/Ireland/D-NVRL-20G44423/2020\|EPI_ISL_60508 | ···················· | ···················· | ···················· | ···················· | ···················· | ···················· | |
| N | 6889 hCoV-19/Ireland/D-NVRL-20G35301/2020\|EPI_ISL_60508 | ···················· | ···················· | ···················· | ···················· | ···················· | ···················· | |
| N | 6890 hCoV-19/Ireland/WW-NVRL-20G31821/2020\|EPI_ISL_6050 | ···················· | ···················· | ···················· | ···················· | ···················· | ···················· | |
| N | 6891 hCoV-19/Ireland/D-NVRL-20G35272/2020\|EPI_ISL_60508 | ···················· | ···················· | ···················· | ···················· | ···················· | ···················· | |
| N | 6892 hCoV-19/Ireland/D-NVRL-20G41351/2020\|EPI_ISL_60508 | ···················· | ···················· | ···················· | ···················· | ···················· | ···················· | |
| N | 6893 hCoV-19/Ireland/D-NVRL-20G44420/2020\|EPI_ISL_60508 | ···················· | ···················· | ···················· | ···················· | ···················· | ···················· | |
| N | 6894 hCoV-19/Ireland/MH-NVRL-20G44535/2020\|EPI_ISL_6050 | ···················· | ···················· | ···················· | ···················· | ···················· | ···················· | |
| N | 6895 hCoV-19/Ireland/D-NVRL-20G44428/2020\|EPI_ISL_60508 | ···················· | ···················· | ···················· | ···················· | ···················· | ···················· | |
| N | 6896 hCoV-19/Ireland/D-NVRL-20G44368/2020\|EPI_ISL_60510 | ···················· | ···················· | ···················· | ···················· | ···················· | ···················· | |
| N | 6897 hCoV-19/Ireland/D-NVRL-20G32263/2020\|EPI_ISL_60510 | ···················· | ···················· | ···················· | ···················· | ···················· | ···················· | |
| N | 6898 hCoV-19/Ireland/D-NVRL-20G35448/2020\|EPI_ISL_60510 | ···················· | ···················· | ···················· | ···················· | ···················· | ···················· | |
| N | 6899 hCoV-19/Ireland/D-NVRL-20G35372/2020\|EPI_ISL_60510 | ···················· | ···················· | ···················· | ···················· | ···················· | ···················· | |
| N | 6900 hCoV-19/Ireland/D-NVRL-20G35404/2020\|EPI_ISL_60510 | ···················· | ···················· | ···················· | ···················· | ···················· | ···················· | |
| N | 6901 hCoV-19/Ireland/D-NVRL-20G35380/2020\|EPI_ISL_60510 | ···················· | ···················· | ···················· | ···················· | ···················· | ···················· | |
| N | 6902 hCoV-19/Ireland/D-NVRL-20G35452/2020\|EPI_ISL_60510 | ···················· | ···················· | ···················· | ···················· | ···················· | ···················· | |
| N | 6903 hCoV-19/Ireland/D-NVRL-20G32271/2020\|EPI_ISL_60510 | ···················· | ···················· | ···················· | ···················· | ···················· | ···················· | |
| N | 6904 hCoV-19/Ireland/OY-NVRL-20G34036/2020\|EPI_ISL_6051 | ···················· | ···················· | ···················· | ···················· | ···················· | ···················· | |
| N | 6905 hCoV-19/Ireland/D-NVRL-20G32281/2020\|EPI_ISL_60511 | ···················· | ···················· | ···················· | ···················· | ···················· | ···················· | |
| N | 6906 hCoV-19/Ireland/WH-NVRL-20G32446/2020\|EPI_ISL_6051 | ···················· | ···················· | ···················· | ···················· | ···················· | ···················· | |
| N | 6907 hCoV-19/Ireland/MH-NVRL-20G32450/2020\|EPI_ISL_6051 | ···················· | ···················· | ···················· | ···················· | ···················· | ···················· | |
| N | 6908 hCoV-19/Ireland/D-NVRL-20G32296/2020\|EPI_ISL_60512 | ···················· | ···················· | ···················· | ···················· | ···················· | ···················· | |
| N | 6909 hCoV-19/Ireland/WW-NVRL-20G35608/2020\|EPI_ISL_6051 | ···················· | ···················· | ···················· | ···················· | ···················· | ···················· | |
| N | 6910 hCoV-19/Ireland/D-NVRL-20G35425/2020\|EPI_ISL_60512 | ···················· | ···················· | ···················· | ···················· | ···················· | ···················· | |
| N | 6911 hCoV-19/Ireland/CO-NVRL-20G32584/2020\|EPI_ISL_6051 | ···················· | ···················· | ···················· | ···················· | ···················· | ···················· | |
| N | 6912 hCoV-19/Ireland/D-NVRL-20G32314/2020\|EPI_ISL_60512 | ···················· | ···················· | ···················· | ···················· | ···················· | ···················· | |
| N | 6913 hCoV-19/Palestine/10/2020\|EPI_ISL_596500\|2020-07-1 | ···················· | ···················· | ···················· | ···················· | ···················· | ···················· | |
| N | 6914 hCoV-19/Palestine/11/2020\|EPI_ISL_596501\|2020-07-1 | ···················· | ···················· | ···················· | ···················· | ···················· | ···················· | |
| N | 6915 hCoV-19/Palestine/13/2020\|EPI_ISL_596502\|2020-03-0 | ···················· | ···················· | ···················· | ···················· | ···················· | ···················· | |
| N | 6916 hCoV-19/Palestine/14/2020\|EPI_ISL_596503\|2020-03-1 | ···················· | ···················· | ···················· | ···················· | ···················· | ···················· | |
| N | 6917 hCoV-19/Palestine/15/2020\|EPI_ISL_596504\|2020-03-0 | ···················· | ···················· | ···················· | ···················· | ···················· | ···················· | |

| | | | | | | | | |
|---|---|---|---|---|---|---|---|---|
| N | 6918 | hCoV-19/Palestine/19/2020\|EPI_ISL_596506\|2020-03-1 | ···················· | ···················· | ···················· | ···················· | ···················· | ···················· |
| N | 6919 | hCoV-19/Palestine/1/2020\|EPI_ISL_596507\|2020-07-15 | ···················· | ···················· | ···················· | ···················· | ···················· | ···················· |
| N | 6920 | hCoV-19/Palestine/21/2020\|EPI_ISL_596508\|2020-03-1 | ···················· | ···················· | ···················· | ···················· | ···················· | ···················· |
| N | 6921 | hCoV-19/Palestine/22/2020\|EPI_ISL_596509\|2020-03-2 | ···················· | ···················· | ···················· | ···················· | ···················· | ···················· |
| N | 6922 | hCoV-19/Palestine/23/2020\|EPI_ISL_596510\|2020-03-2 | ···················· | ···················· | ···················· | ···················· | ···················· | ···················· |
| N | 6923 | hCoV-19/Palestine/24/2020\|EPI_ISL_596511\|2020-03-2 | ···················· | ···················· | ···················· | ···················· | ···················· | ···················· |
| N | 6924 | hCoV-19/Palestine/28/2020\|EPI_ISL_596512\|2020-03-2 | ···················· | ···················· | ···················· | ···················· | ···················· | ···················· |
| N | 6925 | hCoV-19/Palestine/29/2020\|EPI_ISL_596513\|2020-03-3 | ···················· | ···················· | ···················· | ···················· | ···················· | ···················· |
| N | 6926 | hCoV-19/Palestine/2/2020\|EPI_ISL_596514\|2020-07-15 | ···················· | ···················· | ···················· | ···················· | ···················· | ···················· |
| N | 6927 | hCoV-19/Palestine/30/2020\|EPI_ISL_596515\|2020-03-3 | ···················· | ···················· | ···················· | ···················· | ···················· | ···················· |
| N | 6928 | hCoV-19/Palestine/31/2020\|EPI_ISL_596516\|2020-03-3 | ···················· | ···················· | ···················· | ···················· | ···················· | ···················· |
| N | 6929 | hCoV-19/Palestine/32/2020\|EPI_ISL_596517\|2020-03-3 | ···················· | ···················· | ···················· | ···················· | ···················· | ···················· |
| N | 6930 | hCoV-19/Palestine/33/2020\|EPI_ISL_596518\|2020-03-3 | ···················· | ···················· | ···················· | ···················· | ···················· | ···················· |
| N | 6931 | hCoV-19/Palestine/36/2020\|EPI_ISL_596520\|2020-07-2 | ···················· | ···················· | ···················· | ···················· | ···················· | ···················· |
| N | 6932 | hCoV-19/Palestine/66/2020\|EPI_ISL_596543\|2020-06-2 | ···················· | ···················· | ···················· | ···················· | ···················· | ···················· |
| N | 6933 | hCoV-19/Palestine/68/2020\|EPI_ISL_596544\|2020-07-0 | ···················· | ···················· | ···················· | ···················· | ···················· | ···················· |
| N | 6934 | hCoV-19/Palestine/69/2020\|EPI_ISL_596545\|2020-06-2 | ···················· | ···················· | ···················· | ···················· | ···················· | ···················· |
| N | 6935 | hCoV-19/Palestine/71/2020\|EPI_ISL_596547\|2020-07-1 | ···················· | ···················· | ···················· | ···················· | ···················· | ···················· |
| N | 6936 | hCoV-19/Palestine/72/2020\|EPI_ISL_596548\|2020-06-2 | ···················· | ···················· | ···················· | ···················· | ···················· | ···················· |
| N | 6937 | hCoV-19/Palestine/73/2020\|EPI_ISL_596549\|2020-06-2 | ···················· | ···················· | ···················· | ···················· | ···················· | ···················· |
| N | 6938 | hCoV-19/Palestine/74/2020\|EPI_ISL_596550\|2020-06-2 | ···················· | ···················· | ···················· | ···················· | ···················· | ···················· |
| N | 6939 | hCoV-19/Palestine/75/2020\|EPI_ISL_596551\|2020-07-1 | ···················· | ···················· | ···················· | ···················· | ···················· | ···················· |
| N | 6940 | hCoV-19/Palestine/76/2020\|EPI_ISL_596552\|2020-06-2 | ···················· | ···················· | ···················· | ···················· | ···················· | ···················· |
| N | 6941 | hCoV-19/Palestine/77/2020\|EPI_ISL_596553\|2020-06-2 | ···················· | ···················· | ···················· | ···················· | ···················· | ···················· |
| N | 6942 | hCoV-19/Palestine/78/2020\|EPI_ISL_596554\|2020-06-2 | ···················· | ···················· | ···················· | ···················· | ···················· | ···················· |
| N | 6943 | hCoV-19/Palestine/7/2020\|EPI_ISL_596555\|2020-07-17 | ···················· | ···················· | ···················· | ···················· | ···················· | ···················· |
| N | 6944 | hCoV-19/Palestine/80/2020\|EPI_ISL_596556\|2020-07-0 | ···················· | ···················· | ···················· | ···················· | ···················· | ···················· |
| N | 6945 | hCoV-19/Palestine/82/2020\|EPI_ISL_596557\|2020-07-0 | ···················· | ···················· | ···················· | ···················· | ···················· | ···················· |
| N | 6946 | hCoV-19/Palestine/84/2020\|EPI_ISL_596558\|2020-06-2 | ···················· | ····A··············· | ···················· | ···················· | ···················· | ···················· |
| N | 6947 | hCoV-19/Palestine/86/2020\|EPI_ISL_596559\|2020-06-2 | ···················· | ···················· | ···················· | ···················· | ···················· | ···················· |
| N | 6948 | hCoV-19/Palestine/87/2020\|EPI_ISL_596560\|2020-06-2 | ···················· | ····A··············· | ···················· | ···················· | ···················· | ···················· |
| N | 6949 | hCoV-19/Palestine/88/2020\|EPI_ISL_596561\|2020-06-2 | ···················· | ····A··············· | ···················· | ···················· | ···················· | ···················· |
| N | 6950 | hCoV-19/Palestine/89/2020\|EPI_ISL_596562\|2020-06-2 | ···················· | ···················· | ···················· | ···················· | ···················· | ···················· |
| N | 6951 | hCoV-19/Palestine/90/2020\|EPI_ISL_596563\|2020-06-2 | ···················· | ···················· | ···················· | ···················· | ···················· | ···················· |
| N | 6952 | hCoV-19/Palestine/94/2020\|EPI_ISL_596564\|2020-07-1 | ···················· | ···················· | ···················· | ···················· | ···················· | ···················· |
| N | 6953 | hCoV-19/Palestine/95/2020\|EPI_ISL_596565\|2020-07-1 | ···················· | ···················· | ···················· | ···················· | ···················· | ···················· |
| N | 6954 | hCoV-19/Palestine/96/2020\|EPI_ISL_596566\|2020-07 | ···················· | ···················· | ···················· | ···················· | ···················· | ···················· |
| N | 6955 | hCoV-19/Palestine/9/2020\|EPI_ISL_596567\|2020-07-17 | ···················· | ···················· | ···················· | ···················· | ···················· | ···················· |
| N | 6956 | hCoV-19/USA/MI-UM-10035734226/2020\|EPI_ISL_596569\| | ···················· | ·············A······ | ···················· | ···················· | ···················· | ···················· |
| N | 6957 | hCoV-19/USA/MI-UM-10036287673/2020\|EPI_ISL_596570\| | ···················· | ···················· | ···················· | ···················· | ···················· | ···················· |
| N | 6958 | hCoV-19/USA/MI-UM-10036281666/2020\|EPI_ISL_596571\| | ···················· | ···················· | ···················· | ···················· | ···················· | ···················· |
| N | 6959 | hCoV-19/USA/MI-UM-10036223985/2020\|EPI_ISL_596572\| | ···················· | ···················· | ···················· | ···················· | ···················· | ···················· |
| N | 6960 | hCoV-19/USA/MI-UM-10036246523/2020\|EPI_ISL_596574\| | ···················· | ···················· | ···················· | ···················· | ···················· | ···················· |
| N | 6961 | hCoV-19/USA/MI-UM-10036250932/2020\|EPI_ISL_596576\| | ···················· | ·A·················· | ···················· | ···················· | ···················· | ···················· |
| N | 6962 | hCoV-19/USA/MI-UM-10036242970/2020\|EPI_ISL_596577\| | ···················· | ···················· | ···················· | ···················· | ···················· | ···················· |
| N | 6963 | hCoV-19/USA/MI-UM-10036287406/2020\|EPI_ISL_596578\| | ···················· | ···················· | ···················· | ···················· | ···················· | ···················· |
| N | 6964 | hCoV-19/USA/MI-UM-10036249409/2020\|EPI_ISL_596579\| | ···················· | ···················· | ···················· | ···················· | ···················· | ···················· |
| N | 6965 | hCoV-19/USA/MI-UM-10036277128/2020\|EPI_ISL_596580\| | ···················· | ···················· | ···················· | ···················· | ····NN·············· | ···················· |
| N | 6966 | hCoV-19/USA/MI-UM-10036253517/2020\|EPI_ISL_596582\| | ···················· | ···················· | ···················· | ···················· | ···················· | ···················· |
| N | 6967 | hCoV-19/USA/MI-UM-10036286140/2020\|EPI_ISL_596583\| | ···················· | ···················· | ···················· | ···················· | ···················· | ···················· |
| N | 6969 | hCoV-19/USA/MI-UM-10036297467/2020\|EPI_ISL_596585\| | ···················· | ·A·················· | ···················· | ···················· | ···················· | ···················· |
| N | 6971 | hCoV-19/England/ALDP-A501D7/2020\|EPI_ISL_597830\|20 | ···················· | ···················· | ···················· | ···················· | ···················· | ···················· |
| N | 6972 | hCoV-19/England/ALDP-A506F6/2020\|EPI_ISL_597831\|20 | ···················· | ···················· | ···················· | ···················· | ···················· | ··A················· |
| N | 6973 | hCoV-19/England/ALDP-A50984/2020\|EPI_ISL_597832\|20 | ···················· | ···················· | ···················· | ···················· | ···················· | ···················· |
| N | 6974 | hCoV-19/England/ALDP-A51060/2020\|EPI_ISL_597833\|20 | ···················· | ···················· | ···················· | ···················· | ···················· | ···················· |
| N | 6975 | hCoV-19/England/ALDP-A504ED/2020\|EPI_ISL_597834\|20 | ···················· | ···················· | ···················· | ···················· | ···················· | ···················· |
| N | 6976 | hCoV-19/England/ALDP-A4FE4D/2020\|EPI_ISL_597835\|20 | ···················· | ···················· | ···················· | ···················· | ···················· | ···················· |
| N | 6977 | hCoV-19/England/ALDP-A5066F/2020\|EPI_ISL_597836\|20 | ···················· | ···················· | ···················· | ···················· | ···················· | ···················· |
| N | 6978 | hCoV-19/England/ALDP-A507E4/2020\|EPI_ISL_597837\|20 | ···················· | ···················· | ···················· | ···················· | ···················· | ···················· |
| N | 6979 | hCoV-19/England/ALDP-A506BA/2020\|EPI_ISL_597838\|20 | ···················· | ···················· | ···················· | ···················· | ···················· | ···················· |
| N | 6980 | hCoV-19/England/ALDP-A504A1/2020\|EPI_ISL_597839\|20 | ···················· | ···················· | ···················· | ···················· | ···················· | ···················· |
| N | 6981 | hCoV-19/England/ALDP-A50EEF/2020\|EPI_ISL_597841\|20 | ···················· | ···················· | ···················· | ···················· | ···················· | ···················· |
| N | 6982 | hCoV-19/England/ALDP-A50B24/2020\|EPI_ISL_597842\|20 | ···················· | ···················· | ···················· | ···················· | ···················· | ···················· |
| N | 6983 | hCoV-19/England/ALDP-A50B60/2020\|EPI_ISL_597843\|20 | ···················· | ···················· | ···················· | ···················· | ···················· | ···················· |
| N | 6984 | hCoV-19/England/ALDP-A4FC52/2020\|EPI_ISL_597844\|20 | ···················· | ·········NN········· | ···················· | ···················· | ···················· | ···················· |
| N | 6985 | hCoV-19/England/ALDP-A51015/2020\|EPI_ISL_597845\|20 | ···················· | ···················· | ···················· | ···················· | ···················· | ···················· |
| N | 6986 | hCoV-19/England/ALDP-A5017D/2020\|EPI_ISL_597846\|20 | ···················· | ···················· | ···················· | ···················· | ···················· | ···················· |
| N | 6987 | hCoV-19/England/ALDP-A4FF68/2020\|EPI_ISL_597847\|20 | ···················· | ···················· | ···················· | ···················· | ···················· | ···················· |
| N | 6988 | hCoV-19/England/ALDP-A51103/2020\|EPI_ISL_597848\|20 | ···················· | ···················· | ···················· | ···················· | ···················· | ···················· |
| N | 6989 | hCoV-19/England/ALDP-A501E6/2020\|EPI_ISL_597849\|20 | ···················· | ···················· | ···················· | ···················· | ···················· | ···················· |
| N | 6990 | hCoV-19/England/ALDP-A4F9D3/2020\|EPI_ISL_597850\|20 | ···················· | ···················· | ···················· | ···················· | ···················· | ···················· |
| N | 6991 | hCoV-19/England/ALDP-A509B1/2020\|EPI_ISL_597851\|20 | ···················· | ···················· | ···················· | ···················· | ···················· | ···················· |
| N | 6992 | hCoV-19/England/ALDP-A4FD7D/2020\|EPI_ISL_597874\|20 | ···················· | ···················· | ···················· | ···················· | ···················· | ···················· |
| N | 6993 | hCoV-19/England/ALDP-A4FC70/2020\|EPI_ISL_597875\|20 | ···················· | ···················· | ···················· | ···················· | ···················· | ···················· |
| N | 6994 | hCoV-19/England/ALDP-A503B3/2020\|EPI_ISL_597876\|20 | ···················· | ···················· | ···················· | ···················· | ···················· | ···················· |
| N | 6995 | hCoV-19/England/ALDP-A50A36/2020\|EPI_ISL_597877\|20 | ···················· | ···················· | ···················· | ···················· | ···················· | ···················· |
| N | 6996 | hCoV-19/England/ALDP-A500CB/2020\|EPI_ISL_597878\|20 | ···················· | ···················· | ···················· | ···················· | ···················· | ···················· |
| N | 6997 | hCoV-19/England/ALDP-A4FFE0/2020\|EPI_ISL_597881\|20 | ···················· | ···················· | ···················· | ···················· | ···················· | ···················· |
| N | 6998 | hCoV-19/England/ALDP-A501AA/2020\|EPI_ISL_597882\|20 | ···················· | ···················· | ···················· | ···················· | ···················· | ···················· |
| N | 6999 | hCoV-19/England/ALDP-A50939/2020\|EPI_ISL_597883\|20 | ···················· | ···················· | ···················· | ···················· | ···················· | ···················· |

| | | | | | | | | |
|---|---|---|---|---|---|---|---|---|
| N | 7000 | hCoV-19/England/ALDP-A51BD8/2020\|EPI_ISL_597884\|20 | ···················· | ···················· | ····················· | ···················· | ···················· | ···················· |
| N | 7001 | hCoV-19/England/ALDP-A521F3/2020\|EPI_ISL_597885\|20 | ···················· | ···················· | ····················· | ···················· | ···················· | ···················· |
| N | 7002 | hCoV-19/England/ALDP-A51428/2020\|EPI_ISL_597886\|20 | ···················· | ···················· | ····················· | ···················· | ···················· | ···················· |
| N | 7003 | hCoV-19/England/ALDP-A52524/2020\|EPI_ISL_597887\|20 | ···················· | ···················· | ····················· | ···················· | ···················· | ···················· |
| N | 7004 | hCoV-19/England/ALDP-A5172F/2020\|EPI_ISL_597888\|20 | ···················· | ···················· | ····················· | ···················· | ···················· | ···················· |
| N | 7005 | hCoV-19/England/ALDP-A5168C/2020\|EPI_ISL_597889\|20 | ···················· | ···················· | ····················· | ···················· | ···················· | ···················· |
| N | 7006 | hCoV-19/England/ALDP-A51507/2020\|EPI_ISL_597890\|20 | ···················· | ···················· | ····················· | ···················· | ···················· | ···················· |
| N | 7007 | hCoV-19/England/ALDP-A52858/2020\|EPI_ISL_597891\|20 | ···················· | ···················· | ····················· | ···················· | ···················· | ···················· |
| N | 7008 | hCoV-19/England/ALDP-A51FCD/2020\|EPI_ISL_597892\|20 | ···················· | ···················· | ····················· | ···················· | ···················· | ···················· |
| N | 7009 | hCoV-19/England/ALDP-A51437/2020\|EPI_ISL_597893\|20 | ···················· | ···················· | ····················· | ···················· | ···················· | ···················· |
| N | 7010 | hCoV-19/England/ALDP-A52427/2020\|EPI_ISL_597894\|20 | ···················· | ···················· | ····················· | ···················· | ···················· | ···················· |
| N | 7011 | hCoV-19/England/ALDP-A52481/2020\|EPI_ISL_597895\|20 | ···················· | ···················· | ····················· | ···················· | ···················· | ···················· |
| N | 7012 | hCoV-19/England/ALDP-A524EB/2020\|EPI_ISL_597896\|20 | ···················· | ···················· | ····················· | ···················· | ···················· | ···················· |
| N | 7013 | hCoV-19/England/ALDP-A51B50/2020\|EPI_ISL_597897\|20 | ···················· | ···················· | ····················· | ···················· | ···················· | ···················· |
| N | 7014 | hCoV-19/England/ALDP-A5173E/2020\|EPI_ISL_597898\|20 | ···················· | ···················· | ····················· | ···················· | ···················· | ···················· |
| N | 7015 | hCoV-19/England/ALDP-A5220F/2020\|EPI_ISL_597899\|20 | ···················· | ···················· | ····················· | ···················· | ···················· | ···················· |
| N | 7016 | hCoV-19/England/QEUH-9FF4B4/2020\|EPI_ISL_600160\|20 | ···················· | ···················· | ····················· | ···················· | ···················· | ···················· |
| N | 7017 | hCoV-19/England/QEUH-9FF8F4/2020\|EPI_ISL_600161\|20 | ···················· | ·A·················· | ····················· | ···················· | ···················· | ··A················· |
| N | 7018 | hCoV-19/England/QEUH-9FF38A/2020\|EPI_ISL_600162\|20 | ···················· | ···················· | ····················· | ···················· | ···················· | ···················· |
| N | 7019 | hCoV-19/England/QEUH-9FF487/2020\|EPI_ISL_600163\|20 | ···················· | ···················· | ····················· | ···················· | ···················· | ···················· |
| N | 7020 | hCoV-19/England/QEUH-9FFD22/2020\|EPI_ISL_600164\|20 | ···················· | ···················· | ····················· | ···················· | ···················· | ···················· |
| N | 7021 | hCoV-19/England/QEUH-9FFFD1/2020\|EPI_ISL_600165\|20 | ···················· | ···················· | ····················· | ···················· | ···················· | ···················· |
| N | 7022 | hCoV-19/England/QEUH-9FF4C3/2020\|EPI_ISL_600166\|20 | ···················· | ···················· | ····················· | ···················· | ···················· | ···················· |
| N | 7023 | hCoV-19/England/QEUH-9FF4A5/2020\|EPI_ISL_600167\|20 | ···················· | ···················· | ····················· | ···················· | ···················· | ···················· |
| N | 7024 | hCoV-19/England/QEUH-A00437/2020\|EPI_ISL_600168\|20 | ···················· | ···················· | ····················· | ···················· | ···················· | ···················· |
| N | 7025 | hCoV-19/England/QEUH-9FF557/2020\|EPI_ISL_600170\|20 | ···················· | ···················· | ····················· | ···················· | ···················· | ···················· |
| N | 7026 | hCoV-19/Scotland/QEUH-9FFE98/2020\|EPI_ISL_600171\|2 | ···················· | ···················· | ····················· | ···················· | ···················· | ···················· |
| N | 7027 | hCoV-19/England/QEUH-A00103/2020\|EPI_ISL_600173\|20 | ···················· | ···················· | ····················· | ···················· | ···················· | ···················· |
| N | 7028 | hCoV-19/Scotland/QEUH-A00385/2020\|EPI_ISL_600174\|2 | ···················· | ···················· | ····················· | ···················· | ···················· | ···················· |
| N | 7029 | hCoV-19/Scotland/QEUH-9FF760/2020\|EPI_ISL_600175\|2 | ···················· | ···················· | ····················· | ···················· | ···················· | ···················· |
| N | 7030 | hCoV-19/England/QEUH-9FFCF8/2020\|EPI_ISL_600176\|20 | ···················· | ···················· | ····················· | ···················· | ···················· | ···················· |
| N | 7031 | hCoV-19/England/QEUH-9FFC07/2020\|EPI_ISL_600177\|20 | ···················· | ···················· | ····················· | ···················· | ···················· | ···················· |
| N | 7032 | hCoV-19/England/QEUH-9FF5DF/2020\|EPI_ISL_600178\|20 | ···················· | ···················· | ····················· | ···················· | ···················· | ···················· |
| N | 7033 | hCoV-19/England/QEUH-A004BF/2020\|EPI_ISL_600179\|20 | ···················· | ···················· | ····················· | ···················· | ···················· | ···················· |
| N | 7034 | hCoV-19/England/QEUH-A00491/2020\|EPI_ISL_600180\|20 | ···················· | ···················· | ····················· | ···················· | ···················· | ···················· |
| N | 7035 | hCoV-19/Scotland/QEUH-A003A3/2020\|EPI_ISL_600181\|2 | ···················· | ···················· | ····················· | ···················· | ···················· | ···················· |
| N | 7036 | hCoV-19/England/QEUH-9FFAFE/2020\|EPI_ISL_600182\|20 | ···················· | ···················· | ····················· | ···················· | ···················· | ···················· |
| N | 7037 | hCoV-19/England/QEUH-9FF609/2020\|EPI_ISL_600183\|20 | ···················· | ···················· | ····················· | ···················· | ···················· | ···················· |
| N | 7038 | hCoV-19/Scotland/QEUH-9FFEF2/2020\|EPI_ISL_600184\|2 | ···················· | ···················· | ····················· | ···················· | ···················· | ···················· |
| N | 7039 | hCoV-19/England/QEUH-A001E5/2020\|EPI_ISL_600185\|20 | ···················· | ···················· | ····················· | ···················· | ···················· | ···················· |
| N | 7040 | hCoV-19/England/QEUH-A00455/2020\|EPI_ISL_600186\|20 | ···················· | ···················· | ····················· | ···················· | ···················· | ···················· |
| N | 7041 | hCoV-19/England/QEUH-9FF5C0/2020\|EPI_ISL_600187\|20 | ···················· | ···················· | ····················· | ···················· | ···················· | ···················· |
| N | 7042 | hCoV-19/England/QEUH-9FF830/2020\|EPI_ISL_600188\|20 | ···················· | ···················· | ····················· | ···················· | ···················· | ···················· |
| N | 7043 | hCoV-19/England/QEUH-9FF2F6/2020\|EPI_ISL_600189\|20 | ···················· | ···················· | ····················· | ···················· | ···················· | ···················· |
| N | 7044 | hCoV-19/Scotland/QEUH-9FFF2C/2020\|EPI_ISL_600191\|2 | ···················· | ···················· | ····················· | ···················· | ···················· | ···················· |
| N | 7045 | hCoV-19/England/QEUH-A001D6/2020\|EPI_ISL_600192\|20 | ···················· | ···················· | ····················· | ···················· | ···················· | ···················· |
| N | 7046 | hCoV-19/England/QEUH-A001C7/2020\|EPI_ISL_600193\|20 | ···················· | ···················· | ····················· | ···················· | ···················· | ···················· |
| N | 7048 | hCoV-19/England/QEUH-9FF205/2020\|EPI_ISL_600199\|20 | ···················· | ···················· | ····················· | ···················· | ···················· | ···················· |
| N | 7049 | hCoV-19/England/QEUH-9FFC9E/2020\|EPI_ISL_600217\|20 | ···················· | ···················· | ····················· | ···················· | ···················· | ···················· |
| N | 7050 | hCoV-19/England/QEUH-9FFFA4/2020\|EPI_ISL_600220\|20 | ···················· | ···················· | ····················· | ···················· | ···················· | ···················· |
| N | 7051 | hCoV-19/England/QEUH-9FF575/2020\|EPI_ISL_600222\|20 | ···················· | ···················· | ····················· | ···················· | ···················· | ···················· |
| N | 7052 | hCoV-19/England/QEUH-9FFAC1/2020\|EPI_ISL_600223\|20 | ···················· | ···················· | ····················· | ···················· | ···················· | ···················· |
| N | 7053 | hCoV-19/England/QEUH-A00482/2020\|EPI_ISL_600224\|20 | ···················· | ···················· | ····················· | ···················· | ···················· | ···················· |
| N | 7054 | hCoV-19/England/QEUH-9FF469/2020\|EPI_ISL_600225\|20 | ···················· | ···················· | ····················· | ···················· | ···················· | ···················· |
| N | 7055 | hCoV-19/England/QEUH-9FF751/2020\|EPI_ISL_600226\|20 | ···················· | ···················· | ····················· | ···················· | ···················· | ···················· |
| N | 7056 | hCoV-19/England/QEUH-9FF1CC/2020\|EPI_ISL_600227\|20 | ···················· | ···················· | ····················· | ···················· | ···················· | ···················· |
| N | 7057 | hCoV-19/Scotland/QEUH-9FFCAD/2020\|EPI_ISL_600228\|2 | ···················· | ···················· | ····················· | ···················· | ···················· | ···················· |
| N | 7058 | hCoV-19/England/QEUH-9FFB82/2020\|EPI_ISL_600229\|20 | ···················· | ···················· | ····················· | ···················· | ···················· | ···················· |
| N | 7059 | hCoV-19/England/QEUH-A002F1/2020\|EPI_ISL_600230\|20 | ···················· | ···················· | ····················· | ···················· | ···················· | ···················· |
| N | 7060 | hCoV-19/England/QEUH-A0091A/2020\|EPI_ISL_600231\|20 | ···················· | ···················· | ····················· | ···················· | ···················· | ···················· |
| N | 7061 | hCoV-19/England/QEUH-A00473/2020\|EPI_ISL_600232\|20 | ···················· | ···················· | ····················· | ···················· | ···················· | ···················· |
| N | 7062 | hCoV-19/England/QEUH-9FF636/2020\|EPI_ISL_600234\|20 | ···················· | ·A·················· | ····················· | ···················· | ···················· | ···················· |
| N | 7063 | hCoV-19/England/QEUH-9FFF95/2020\|EPI_ISL_600236\|20 | ···················· | ···················· | ····················· | ···················· | ···················· | ···················· |
| N | 7064 | hCoV-19/England/QEUH-A004DD/2020\|EPI_ISL_600238\|20 | ···················· | ···················· | ····················· | ···················· | ···················· | ···················· |
| N | 7065 | hCoV-19/England/MILK-9D3CCD/2020\|EPI_ISL_601490\|20 | ···················· | ···················· | ····················· | ···················· | ···················· | ···················· |
| N | 7066 | hCoV-19/England/MILK-9D4840/2020\|EPI_ISL_601491\|20 | ···················· | ···················· | ····················· | ···················· | ···················· | ···················· |
| N | 7067 | hCoV-19/England/MILK-9D50ED/2020\|EPI_ISL_601492\|20 | ···················· | ···················· | ····················· | ···················· | ···················· | ···················· |
| N | 7068 | hCoV-19/England/CAMC-9D2174/2020\|EPI_ISL_601493\|20 | ···················· | ···················· | ····················· | ···················· | ···················· | ···················· |
| N | 7069 | hCoV-19/England/MILK-9D3ED6/2020\|EPI_ISL_601494\|20 | ···················· | ···················· | ····················· | ···················· | ···················· | ···················· |
| N | 7070 | hCoV-19/England/CAMC-9D203B/2020\|EPI_ISL_601495\|20 | ···················· | ···················· | ····················· | ···················· | ···················· | ···················· |
| N | 7071 | hCoV-19/England/MILK-9D400C/2020\|EPI_ISL_601496\|20 | ···················· | ···················· | ····················· | ···················· | ···················· | ···················· |
| N | 7072 | hCoV-19/Wales/MILK-9D495C/2020\|EPI_ISL_601497\|2020 | ···················· | ···················· | ····················· | ···················· | ···················· | ···················· |
| N | 7073 | hCoV-19/England/MILK-9D4172/2020\|EPI_ISL_601498\|20 | ···················· | ···················· | ····················· | ···················· | ···················· | ···················· |
| N | 7074 | hCoV-19/England/CAMC-9D1D62/2020\|EPI_ISL_601499\|20 | ···················· | ···················· | ····················· | ···················· | ···················· | ···················· |
| N | 7075 | hCoV-19/England/MILK-9D3A4B/2020\|EPI_ISL_601500\|20 | ···················· | ···················· | ····················· | ···················· | ···················· | ···················· |
| N | 7076 | hCoV-19/England/CAMC-9D1FA8/2020\|EPI_ISL_601501\|20 | ···················· | ···················· | ····················· | ···················· | ···················· | ···················· |
| N | 7077 | hCoV-19/England/MILK-9D4A0E/2020\|EPI_ISL_601502\|20 | ···················· | ···················· | ····················· | ···················· | ···················· | ···················· |
| N | 7078 | hCoV-19/Wales/MILK-9D4D41/2020\|EPI_ISL_601503\|2020 | ···················· | ···················· | ····················· | ···················· | ···················· | ···················· |
| N | 7079 | hCoV-19/Wales/MILK-9D3A3C/2020\|EPI_ISL_601504\|2020 | ···················· | ···················· | ····················· | ···················· | ···················· | ···················· |
| N | 7080 | hCoV-19/England/MILK-9D4743/2020\|EPI_ISL_601505\|20 | ···················· | ···················· | ····················· | ···················· | ···················· | ···················· |

| | | | | | | | | |
|---|---|---|---|---|---|---|---|---|
| N | 7081 | hCoV-19/England/CAMC-9D2253/2020\|EPI_ISL_601506\|20 | ···················· | ···················· | ····················· | ···················· | ···················· | ···················· |
| N | 7082 | hCoV-19/England/MILK-9D40EE/2020\|EPI_ISL_601508\|20 | ···················· | ···················· | ····················· | ···················· | ···················· | ···················· |
| N | 7083 | hCoV-19/Wales/MILK-9D3F6A/2020\|EPI_ISL_601509\|2020 | ···················· | ···················· | ····················· | ···················· | ···················· | ···················· |
| N | 7084 | hCoV-19/Wales/MILK-9D3C36/2020\|EPI_ISL_601510\|2020 | ···················· | ···················· | ····················· | ···················· | ···················· | ···················· |
| N | 7085 | hCoV-19/England/MILK-9D4E02/2020\|EPI_ISL_601511\|20 | ···················· | ···················· | ····················· | ···················· | ···················· | ···················· |
| N | 7086 | hCoV-19/Wales/MILK-9D3C45/2020\|EPI_ISL_601512\|2020 | ···················· | ···················· | ····················· | ···················· | ···················· | ···················· |
| N | 7087 | hCoV-19/England/MILK-9D4682/2020\|EPI_ISL_601513\|20 | ···················· | ···················· | ····················· | ···················· | ···················· | ···················· |
| N | 7088 | hCoV-19/England/MILK-9D4AD1/2020\|EPI_ISL_601514\|20 | ···················· | ····A··············· | ····················· | ···················· | ···················· | ···················· |
| N | 7089 | hCoV-19/England/CAMC-9D1D44/2020\|EPI_ISL_601515\|20 | ···················· | ···················· | ····················· | ···················· | ···················· | ···················· |
| N | 7090 | hCoV-19/Wales/MILK-9D4989/2020\|EPI_ISL_601516\|2020 | ···················· | ·A·················· | ····················· | ···················· | ···················· | ···················· |
| N | 7091 | hCoV-19/Wales/MILK-9D4FE1/2020\|EPI_ISL_601517\|2020 | ···················· | ···················· | ····················· | ···················· | ···················· | ···················· |
| N | 7092 | hCoV-19/England/MILK-9D4BB0/2020\|EPI_ISL_601518\|20 | ···················· | ···················· | ····················· | ···················· | ···················· | ···················· |
| N | 7093 | hCoV-19/Wales/MILK-9D4D32/2020\|EPI_ISL_601519\|2020 | ···················· | ···················· | ····················· | ···················· | ···················· | ···················· |
| N | 7094 | hCoV-19/England/MILK-9D47DA/2020\|EPI_ISL_601520\|20 | ···················· | ···················· | ····················· | ···················· | ···················· | ···················· |
| N | 7095 | hCoV-19/England/MILK-9D46EC/2020\|EPI_ISL_601521\|20 | ···················· | ···················· | ····················· | ···················· | ···················· | ···················· |
| N | 7096 | hCoV-19/England/CAMC-9D2280/2020\|EPI_ISL_601522\|20 | ···················· | ···················· | ····················· | ···················· | ···················· | ···················· |
| N | 7097 | hCoV-19/England/MILK-9D4F0F/2020\|EPI_ISL_601523\|20 | ···················· | ···················· | ····················· | ···················· | ···················· | ···················· |
| N | 7098 | hCoV-19/England/MILK-9D497A/2020\|EPI_ISL_601524\|20 | ···················· | ···················· | ····················· | ···················· | ···················· | ···················· |
| N | 7099 | hCoV-19/England/CAMC-9D1DBD/2020\|EPI_ISL_601550\|20 | ···················· | ···················· | ····················· | ···················· | ···················· | ···················· |
| N | 7100 | hCoV-19/Wales/MILK-9D3BB1/2020\|EPI_ISL_601551\|2020 | ···················· | ···················· | ····················· | ···················· | ···················· | ···················· |
| N | 7101 | hCoV-19/England/MILK-9D4F5A/2020\|EPI_ISL_601552\|20 | ···················· | ···················· | ····················· | ···················· | ···················· | ···················· |
| N | 7102 | hCoV-19/Wales/MILK-9D45C1/2020\|EPI_ISL_601553\|2020 | ···················· | ···················· | ····················· | ···················· | ···················· | ···················· |
| N | 7103 | hCoV-19/England/MILK-9D4A68/2020\|EPI_ISL_601554\|20 | ···················· | ···················· | ····················· | ···················· | ···················· | ···················· |
| N | 7104 | hCoV-19/England/CAMC-9D2235/2020\|EPI_ISL_601555\|20 | ···················· | ···················· | ····················· | ···················· | ···················· | ···················· |
| N | 7105 | hCoV-19/England/MILK-9D46DD/2020\|EPI_ISL_601556\|20 | ···················· | ···················· | ····················· | ···················· | ···················· | ···················· |
| N | 7106 | hCoV-19/England/MILK-9D4AE0/2020\|EPI_ISL_601557\|20 | ···················· | ···················· | ····················· | ···················· | ···················· | ···················· |
| N | 7107 | hCoV-19/England/MILK-9D3D9D/2020\|EPI_ISL_601559\|20 | ···················· | ···················· | ····················· | ···················· | ···················· | ···················· |
| N | 7108 | hCoV-19/England/MILK-9D4057/2020\|EPI_ISL_601560\|20 | ···················· | ···················· | ····················· | ···················· | ···················· | ···················· |
| N | 7109 | hCoV-19/England/MILK-9D4F78/2020\|EPI_ISL_601561\|20 | ···················· | ···················· | ····················· | ···················· | ···················· | ···················· |
| N | 7110 | hCoV-19/England/MILK-9D4B38/2020\|EPI_ISL_601562\|20 | ···················· | ···················· | ····················· | ···················· | ···················· | ···················· |
| N | 7111 | hCoV-19/England/MILK-9D492F/2020\|EPI_ISL_601563\|20 | ···················· | ·A·················· | ····················· | ···················· | ···················· | ···A················ |
| N | 7112 | hCoV-19/England/MILK-9D3E4F/2020\|EPI_ISL_601564\|20 | ···················· | ···················· | ····················· | ···················· | ···················· | ···················· |
| N | 7113 | hCoV-19/Wales/MILK-9D50B0/2020\|EPI_ISL_601565\|2020 | ···················· | ···················· | ····················· | ···················· | ···················· | ···················· |
| N | 7114 | hCoV-19/Wales/MILK-9D4C80/2020\|EPI_ISL_601566\|2020 | ···················· | ···················· | ····················· | ···················· | ···················· | ···················· |
| N | 7115 | hCoV-19/England/CAMC-9D1DDB/2020\|EPI_ISL_601567\|20 | ···················· | ···················· | ····················· | ···················· | ···················· | ···················· |
| N | 7116 | hCoV-19/England/MILK-9D3F5B/2020\|EPI_ISL_601568\|20 | ···················· | ·A·················· | ····················· | ···················· | ···················· | ···A················ |
| N | 7117 | hCoV-19/England/CAMC-9D1E8D/2020\|EPI_ISL_601569\|20 | ···················· | ···················· | ····················· | ···················· | ······T············· | ···················· |
| N | 7118 | hCoV-19/South Africa/KRISP-K003481/2020\|EPI_ISL_60 | ···················· | ···················· | ····················· | ···················· | ···················· | ···················· |
| N | 7119 | hCoV-19/South Africa/KRISP-K003482/2020\|EPI_ISL_60 | ···················· | ···················· | ····················· | ···················· | ···················· | ···················· |
| N | 7120 | hCoV-19/South Africa/KRISP-K003483/2020\|EPI_ISL_60 | ···················· | ···················· | ····················· | ···················· | ···················· | ···················· |
| N | 7121 | hCoV-19/South Africa/KRISP-K003485/2020\|EPI_ISL_60 | ···················· | ···················· | ····················· | ···················· | ···················· | ···················· |
| N | 7122 | hCoV-19/South Africa/KRISP-K003487/2020\|EPI_ISL_60 | ···················· | ···················· | ····················· | ···················· | ···················· | ···················· |
| N | 7123 | hCoV-19/South Africa/KRISP-K003490/2020\|EPI_ISL_60 | ···················· | ···················· | ····················· | ···················· | ···················· | ···················· |
| N | 7124 | hCoV-19/South Africa/KRISP-K003491/2020\|EPI_ISL_60 | ···················· | ···················· | ····················· | ···················· | ···················· | ···················· |
| N | 7125 | hCoV-19/South Africa/KRISP-K003492/2020\|EPI_ISL_60 | ···················· | ···················· | ····················· | ···················· | ···················· | ···················· |
| N | 7126 | hCoV-19/South Africa/KRISP-K003500/2020\|EPI_ISL_60 | ···················· | ···················· | ····················· | ···················· | ···················· | ···················· |
| N | 7128 | hCoV-19/South Africa/KRISP-K003502/2020\|EPI_ISL_60 | ···················· | ···················· | ····················· | ···················· | ···················· | ···················· |
| N | 7130 | hCoV-19/South Africa/KRISP-K003508/2020\|EPI_ISL_60 | ···················· | ···················· | ····················· | ···················· | ···················· | ···················· |
| N | 7131 | hCoV-19/South Africa/KRISP-K003509/2020\|EPI_ISL_60 | ···················· | ···················· | ····················· | ···················· | ···················· | ···················· |
| N | 7132 | hCoV-19/South Africa/KRISP-K003511/2020\|EPI_ISL_60 | ···················· | ···················· | ····················· | ···················· | ···················· | ···················· |
| N | 7133 | hCoV-19/South Africa/KRISP-K003513/2020\|EPI_ISL_60 | ···················· | ···················· | ····················· | ···················· | ···················· | ···················· |
| N | 7134 | hCoV-19/South Africa/KRISP-K003514/2020\|EPI_ISL_60 | ···················· | ···················· | ····················· | ···················· | ···················· | ···················· |
| N | 7135 | hCoV-19/South Africa/KRISP-K003515/2020\|EPI_ISL_60 | ···················· | ···················· | ····················· | ···················· | ···················· | ···················· |
| N | 7136 | hCoV-19/South Africa/KRISP-K003518/2020\|EPI_ISL_60 | ···················· | ···················· | ····················· | ···················· | ···················· | ···················· |
| N | 7137 | hCoV-19/South Africa/KRISP-K003519/2020\|EPI_ISL_60 | ···················· | ·A·················· | ····················· | ···················· | ···················· | ···················· |
| N | 7138 | hCoV-19/South Africa/KRISP-K003520/2020\|EPI_ISL_60 | ···················· | ···················· | ····················· | ···················· | ···················· | ···················· |
| N | 7139 | hCoV-19/South Africa/KRISP-K003521/2020\|EPI_ISL_60 | ···················· | ···················· | ····················· | ···················· | ···················· | ···················· |
| N | 7140 | hCoV-19/South Africa/KRISP-K003522/2020\|EPI_ISL_60 | ···················· | ·A·················· | ····················· | ···················· | ···················· | ···················· |
| N | 7141 | hCoV-19/South Africa/KRISP-K003523/2020\|EPI_ISL_60 | ···················· | ···················· | ····················· | ···················· | ···················· | ···················· |
| N | 7142 | hCoV-19/South Africa/KRISP-K003525/2020\|EPI_ISL_60 | ···················· | ···················· | ····················· | ···················· | ···················· | ···················· |
| N | 7143 | hCoV-19/South Africa/KRISP-K003526/2020\|EPI_ISL_60 | ···················· | ···················· | ····················· | ···················· | ···················· | ···················· |
| N | 7144 | hCoV-19/South Africa/KRISP-K003527/2020\|EPI_ISL_60 | ···················· | ···················· | ····················· | ···················· | ···················· | ···················· |
| N | 7145 | hCoV-19/South Africa/KRISP-K003530/2020\|EPI_ISL_60 | ···················· | ···················· | ····················· | ···················· | ···················· | ···················· |
| N | 7146 | hCoV-19/South Africa/KRISP-K003531/2020\|EPI_ISL_60 | ···················· | ···················· | ····················· | ···················· | ···················· | ···················· |
| N | 7147 | hCoV-19/South Africa/KRISP-K003532/2020\|EPI_ISL_60 | ···················· | ···················· | ····················· | ···················· | ···················· | ···················· |
| N | 7148 | hCoV-19/South Africa/KRISP-K003533/2020\|EPI_ISL_60 | ···················· | ···················· | ····················· | ···················· | ···················· | ···················· |
| N | 7149 | hCoV-19/South Africa/KRISP-K003536/2020\|EPI_ISL_60 | ···················· | ···················· | ····················· | ···················· | ···················· | ···················· |
| N | 7150 | hCoV-19/South Africa/KRISP-K003537/2020\|EPI_ISL_60 | ···················· | ···················· | ····················· | ···················· | ···················· | ···················· |
| N | 7151 | hCoV-19/South Africa/KRISP-K003538/2020\|EPI_ISL_60 | ···················· | ···················· | ····················· | ···················· | ···················· | ···················· |
| N | 7152 | hCoV-19/South Africa/KRISP-K003699/2020\|EPI_ISL_60 | ···················· | ···················· | ····················· | ···················· | ···················· | ···················· |
| N | 7154 | hCoV-19/South Africa/KRISP-K003701/2020\|EPI_ISL_60 | ···················· | ·A·················· | ····················· | ···················· | ···················· | ···················· |
| N | 7155 | hCoV-19/South Africa/KRISP-K003703/2020\|EPI_ISL_60 | ···················· | ···················· | ····················· | ···················· | ···················· | ···················· |
| N | 7156 | hCoV-19/South Africa/KRISP-K003704/2020\|EPI_ISL_60 | ···················· | ···················· | ····················· | ···················· | ···················· | ···················· |
| N | 7157 | hCoV-19/South Africa/KRISP-K003707/2020\|EPI_ISL_60 | ···················· | ···················· | ····················· | ···················· | ···················· | ···················· |
| N | 7158 | hCoV-19/South Africa/KRISP-K003710/2020\|EPI_ISL_60 | ···················· | ···················· | ····················· | ···················· | ···················· | ···················· |
| N | 7159 | hCoV-19/South Africa/KRISP-K003712/2020\|EPI_ISL_60 | ···················· | ···················· | ····················· | ···················· | ···················· | ···················· |
| N | 7160 | hCoV-19/South Africa/KRISP-K003714/2020\|EPI_ISL_60 | ···················· | ···················· | ····················· | ···················· | ···················· | ···················· |
| N | 7162 | hCoV-19/South Africa/KRISP-K003716/2020\|EPI_ISL_60 | ···················· | ···················· | ····················· | ···················· | ···················· | ···················· |
| N | 7163 | hCoV-19/South Africa/KRISP-K003719/2020\|EPI_ISL_60 | ···················· | ···················· | ····················· | ···················· | ···················· | ···················· |
| N | 7164 | hCoV-19/Ireland/D-NVRL-20G35265/2020\|EPI_ISL_60509 | ···················· | ···················· | ····················· | ···················· | ···················· | ···················· |

| | | | | | | | |
|---|---|---|---|---|---|---|---|
| N | 7165 hCoV-19/Ireland/KY-NVRL-20G44380/2020\|EPI_ISL_6050 | .................... | .................... | ..................... | .................... | .................... | .................... |
| N | 7166 hCoV-19/Ireland/D-NVRL-20G35221/2020\|EPI_ISL_60509 | .................... | .................... | ..................... | .................... | .................... | .................... |
| N | 7167 hCoV-19/Ireland/D-NVRL-20G44603/2020\|EPI_ISL_60509 | .................... | .................... | ..................... | .................... | .................... | .................... |
| N | 7168 hCoV-19/Ireland/D-NVRL-20G44441/2020\|EPI_ISL_60509 | .................... | .................... | ..................... | .................... | .................... | .................... |
| N | 7169 hCoV-19/USA/WA-S2869/2020\|EPI_ISL_596400\|2020-09-2 | .................... | .................... | ..................... | .................... | .................... | .................... |
| N | 7170 hCoV-19/USA/WA-S2871/2020\|EPI_ISL_596402\|2020-09-2 | .................... | .................... | ..................... | .................... | .................... | .................... |
| N | 7172 hCoV-19/USA/WA-S2874/2020\|EPI_ISL_596405\|2020-09-2 | .................... | .................... | ..................... | .................... | .................... | .................... |
| N | 7173 hCoV-19/USA/WA-S2875/2020\|EPI_ISL_596406\|2020-09-2 | .................... | .................... | ..................... | .................... | .................... | .................... |
| N | 7174 hCoV-19/USA/WA-S2876/2020\|EPI_ISL_596407\|2020-09-2 | .................... | .A.................. | ..................... | .................... | .................... | .................... |
| N | 7175 hCoV-19/USA/WA-S2879/2020\|EPI_ISL_596410\|2020-09-2 | .................... | .................... | ..................... | .................... | .................... | .................... |
| N | 7176 hCoV-19/USA/WA-S2884/2020\|EPI_ISL_596415\|2020-09-2 | .................... | .A.................. | ..................... | .................... | .................... | .................... |
| N | 7177 hCoV-19/USA/WA-S2886/2020\|EPI_ISL_596417\|2020-09-2 | .................... | .................... | ..................... | .................... | .................... | .................... |
| N | 7178 hCoV-19/USA/WA-S2887/2020\|EPI_ISL_596418\|2020-09-2 | .................... | .................... | ..................... | .................... | .................... | .................... |
| N | 7179 hCoV-19/USA/WA-S2888/2020\|EPI_ISL_596419\|2020-10-0 | .................... | .................... | ..................... | .................... | .................... | .................... |
| N | 7180 hCoV-19/USA/WA-S2889/2020\|EPI_ISL_596420\|2020-10-0 | .................... | .................... | ..................... | .................... | .................... | .................... |
| N | 7181 hCoV-19/Singapore/1076/2020\|EPI_ISL_596466\|2020-10 | .................... | .................... | ..................... | .................... | .................... | .................... |
| N | 7182 hCoV-19/Singapore/1077/2020\|EPI_ISL_596467\|2020-10 | .................... | .................... | ..................... | .................... | .................... | .................... |
| N | 7183 hCoV-19/Singapore/1078/2020\|EPI_ISL_596468\|2020-09 | .................... | .................... | ..................... | .................... | .................... | .................... |
| N | 7184 hCoV-19/Singapore/1079/2020\|EPI_ISL_596469\|2020-10 | .................... | .................... | ..................... | .................... | .................... | .................... |
| N | 7185 hCoV-19/Singapore/1080/2020\|EPI_ISL_596470\|2020-09 | .................... | .A.................. | ..................... | .................... | .................... | ..A................. |
| N | 7186 hCoV-19/Singapore/1081/2020\|EPI_ISL_596471\|2020-10 | .................... | .................... | ..................... | .................... | .................... | .................... |
| N | 7187 hCoV-19/Singapore/1082/2020\|EPI_ISL_596472\|2020-04 | .................... | .................... | ..................... | .................... | .................... | .................... |
| N | 7188 hCoV-19/Singapore/1083/2020\|EPI_ISL_596473\|2020-10 | .................... | .................... | ..................... | .................... | .................... | .................... |
| N | 7189 hCoV-19/Singapore/1084/2020\|EPI_ISL_596474\|2020-09 | .................... | .A.................. | ..................... | .................... | .................... | .................... |
| N | 7190 hCoV-19/Singapore/1085/2020\|EPI_ISL_596475\|2020-09 | .................... | .A.................. | ..................... | .................... | .................... | .................... |
| N | 7191 hCoV-19/Singapore/1086/2020\|EPI_ISL_596476\|2020-08 | .................... | .................... | ..................... | .................... | .................... | .................... |
| N | 7192 hCoV-19/Singapore/1087/2020\|EPI_ISL_596477\|2020-09 | .................... | .A.................. | ..................... | .................... | .................... | .................... |
| N | 7193 hCoV-19/Singapore/1088/2020\|EPI_ISL_596478\|2020-10 | .................... | .................... | ..................... | .................... | .................... | .................... |
| N | 7194 hCoV-19/Singapore/1089/2020\|EPI_ISL_596479\|2020-09 | .................... | .................... | ..................... | .................... | .................... | .................... |
| N | 7195 hCoV-19/Singapore/1090/2020\|EPI_ISL_596480\|2020-10 | .................... | .................... | ..................... | .................... | .................... | .................... |
| N | 7196 hCoV-19/Singapore/1091/2020\|EPI_ISL_596481\|2020-09 | .................... | .................... | ..................... | .................... | .................... | .................... |
| N | 7197 hCoV-19/Singapore/1092/2020\|EPI_ISL_596482\|2020-10 | .................... | .................... | ..................... | .................... | .................... | .................... |
| N | 7198 hCoV-19/Singapore/1093/2020\|EPI_ISL_596483\|2020-10 | .................... | .................... | ..................... | .................... | .................... | .................... |
| N | 7199 hCoV-19/Singapore/1095/2020\|EPI_ISL_596485\|2020-10 | .................... | .A.................. | ..................... | .................... | .................... | .................... |
| N | 7200 hCoV-19/Singapore/1096/2020\|EPI_ISL_596486\|2020-10 | .................... | .A.................. | ..................... | .................... | .................... | ..A................. |
| N | 7201 hCoV-19/Singapore/1097/2020\|EPI_ISL_596487\|2020-10 | .................... | .................... | ..................... | .................... | .................... | .................... |
| N | 7202 hCoV-19/Singapore/1098/2020\|EPI_ISL_596488\|2020-10 | .................... | .A.................. | ..................... | .................... | .................... | ..A................. |
| N | 7203 hCoV-19/Singapore/1099/2020\|EPI_ISL_596489\|2020-10 | .................... | .................... | ..................... | .................... | .................... | .................... |
| N | 7204 hCoV-19/Singapore/1100/2020\|EPI_ISL_596490\|2020-10 | .................... | .................... | ..................... | .................... | .................... | ..A................. |
| N | 7205 hCoV-19/Singapore/1101/2020\|EPI_ISL_596491\|2020-10 | .................... | .................... | ..................... | .................... | .................... | .................... |
| N | 7206 hCoV-19/Singapore/1102/2020\|EPI_ISL_596492\|2020-10 | .................... | .................... | ..................... | .................... | .................... | .................... |
| N | 7207 hCoV-19/Singapore/1103/2020\|EPI_ISL_596493\|2020-10 | .................... | .A.................. | ..................... | .................... | .................... | ..A................. |
| N | 7208 hCoV-19/Singapore/1104/2020\|EPI_ISL_596494\|2020-10 | .................... | .................... | ..................... | .................... | .................... | .................... |
| N | 7209 hCoV-19/Singapore/1105/2020\|EPI_ISL_596495\|2020-10 | .................... | .................... | ..................... | .................... | .................... | .................... |
| N | 7210 hCoV-19/Singapore/1106/2020\|EPI_ISL_596496\|2020-10 | .................... | .................... | ..................... | .................... | .................... | .................... |
| N | 7211 hCoV-19/England/ALDP-A50368/2020\|EPI_ISL_597700\|20 | .................... | .................... | ..................... | .................... | .................... | .................... |
| N | 7212 hCoV-19/England/ALDP-A50720/2020\|EPI_ISL_597701\|20 | .................... | .................... | ..................... | .................... | .................... | .................... |
| N | 7213 hCoV-19/England/ALDP-A4FB55/2020\|EPI_ISL_597702\|20 | .................... | .................... | ..................... | .................... | .................... | .................... |
| N | 7214 hCoV-19/England/ALDP-A509C0/2020\|EPI_ISL_597703\|20 | .................... | .................... | ..................... | .................... | .................... | .................... |
| N | 7215 hCoV-19/England/ALDP-A51024/2020\|EPI_ISL_597704\|20 | .................... | .................... | ..................... | .................... | .................... | .................... |
| N | 7216 hCoV-19/England/ALDP-A4FFC2/2020\|EPI_ISL_597705\|20 | .................... | .................... | ..................... | .................... | .................... | .................... |
| N | 7217 hCoV-19/England/ALDP-A50C5E/2020\|EPI_ISL_597706\|20 | .................... | .................... | ..................... | .................... | .................... | .................... |
| N | 7218 hCoV-19/England/ALDP-A50B51/2020\|EPI_ISL_597707\|20 | .................... | .................... | ..................... | .................... | .................... | .................... |
| N | 7219 hCoV-19/England/ALDP-A508B4/2020\|EPI_ISL_597708\|20 | .................... | .................... | ..................... | .................... | .................... | .................... |
| N | 7220 hCoV-19/England/ALDP-A4FBFB/2020\|EPI_ISL_597709\|20 | .................... | .................... | ..................... | .................... | .................... | .................... |
| N | 7221 hCoV-19/England/ALDP-A4F9A6/2020\|EPI_ISL_597717\|20 | .................... | .A.................. | ..................... | .................... | .................... | .................... |
| N | 7222 hCoV-19/England/ALDP-A4FA2B/2020\|EPI_ISL_597718\|20 | .................... | .................... | ..................... | .................... | .................... | .................... |
| N | 7223 hCoV-19/England/ALDP-A4FF86/2020\|EPI_ISL_597719\|20 | .................... | .................... | ..................... | .................... | .................... | .................... |
| N | 7224 hCoV-19/England/ALDP-A505DB/2020\|EPI_ISL_597731\|20 | .................... | .................... | ..................... | .................... | .................... | .................... |
| N | 7225 hCoV-19/England/ALDP-A500AD/2020\|EPI_ISL_597732\|20 | .................... | .................... | ..................... | .................... | .................... | .................... |
| N | 7226 hCoV-19/England/ALDP-A5040B/2020\|EPI_ISL_597733\|20 | .................... | .................... | ..................... | .................... | .................... | .................... |
| N | 7227 hCoV-19/England/ALDP-A50799/2020\|EPI_ISL_597734\|20 | .................... | .................... | ..................... | .................... | .................... | .................... |
| N | 7228 hCoV-19/England/ALDP-A50CF4/2020\|EPI_ISL_597735\|20 | .................... | .................... | ..................... | .................... | .................... | .................... |
| N | 7229 hCoV-19/England/ALDP-A50BF7/2020\|EPI_ISL_597736\|20 | .................... | .................... | ..................... | .................... | .................... | .................... |
| N | 7230 hCoV-19/England/ALDP-A50C8B/2020\|EPI_ISL_597737\|20 | .................... | .................... | ..................... | .................... | .................... | .................... |
| N | 7231 hCoV-19/England/ALDP-A506AB/2020\|EPI_ISL_597738\|20 | .................... | .................... | ..................... | .................... | .................... | .................... |
| N | 7232 hCoV-19/England/ALDP-A4FE2F/2020\|EPI_ISL_597739\|20 | .................... | .................... | ..................... | .................... | .................... | .................... |
| N | 7233 hCoV-19/England/ALDP-A4FD9B/2020\|EPI_ISL_597740\|20 | .................... | .................... | ..................... | .................... | .................... | .................... |
| N | 7234 hCoV-19/England/ALDP-A4FA1C/2020\|EPI_ISL_597741\|20 | .................... | .................... | ..................... | .................... | .................... | .................... |
| N | 7235 hCoV-19/England/ALDP-A50F46/2020\|EPI_ISL_597742\|20 | .................... | .................... | ..................... | .................... | .................... | .................... |
| N | 7236 hCoV-19/England/ALDP-A50FBF/2020\|EPI_ISL_597743\|20 | .................... | .A.................. | ..................... | .................... | .................... | ..A................. |
| N | 7237 hCoV-19/England/ALDP-A4FF77/2020\|EPI_ISL_597744\|20 | .................... | .................... | ..................... | .................... | .................... | .................... |
| N | 7238 hCoV-19/England/ALDP-A50ED0/2020\|EPI_ISL_597745\|20 | .................... | .................... | ..................... | .................... | .................... | .................... |
| N | 7239 hCoV-19/England/ALDP-A4FAB2/2020\|EPI_ISL_597746\|20 | .................... | .................... | ..................... | .................... | .................... | .................... |
| N | 7240 hCoV-19/England/ALDP-A500F8/2020\|EPI_ISL_597747\|20 | .................... | .................... | ..................... | .................... | .................... | .................... |
| N | 7241 hCoV-19/England/ALDP-A50975/2020\|EPI_ISL_597748\|20 | .................... | .................... | ..................... | .................... | .................... | .................... |
| N | 7242 hCoV-19/England/ALDP-A50E0D/2020\|EPI_ISL_597749\|20 | .................... | .................... | ..................... | .................... | .................... | .................... |
| N | 7243 hCoV-19/England/ALDP-A50C03/2020\|EPI_ISL_597750\|20 | .................... | .A.................. | ..................... | .................... | .................... | ..A................. |
| N | 7244 hCoV-19/England/ALDP-A4FF0E/2020\|EPI_ISL_597752\|20 | .................... | .................... | ...........Y......... | .................... | .................... | .................... |
| N | 7245 hCoV-19/England/ALDP-A4FCF8/2020\|EPI_ISL_597797\|20 | .................... | .A.................. | ..................... | .................... | .................... | ..A................. |

| | | | | | | | | |
|---|---|---|---|---|---|---|---|---|
| N | 7246 | hCoV-19/England/ALDP-A5059F/2020\|EPI_ISL_597798\|20 | ···················· | ···················· | ···················· | ···················· | ···················· | ···················· |
| N | 7247 | hCoV-19/England/ALDP-A50F82/2020\|EPI_ISL_597799\|20 | ···················· | ···················· | ···················· | ···················· | ···················· | ···················· |
| N | 7248 | hCoV-19/England/MILK-A06668/2020\|EPI_ISL_600100\|20 | ···················· | ···················· | ···················· | ···················· | ···················· | ···················· |
| N | 7249 | hCoV-19/England/MILK-A0620A/2020\|EPI_ISL_600101\|20 | ···················· | ···················· | ···················· | ···················· | ···················· | ···················· |
| N | 7250 | hCoV-19/England/MILK-A05F6E/2020\|EPI_ISL_600102\|20 | ···················· | ···················· | ···················· | ···················· | ···················· | ···················· |
| N | 7251 | hCoV-19/England/MILK-A06194/2020\|EPI_ISL_600105\|20 | ···················· | ···················· | ···················· | ···················· | ···················· | ···················· |
| N | 7252 | hCoV-19/England/MILK-A05F7D/2020\|EPI_ISL_600106\|20 | ···················· | ···················· | ···················· | ···················· | ···················· | ···················· |
| N | 7253 | hCoV-19/England/MILK-A066FF/2020\|EPI_ISL_600118\|20 | ···················· | ···················· | ···················· | ···················· | ···················· | ···················· |
| N | 7254 | hCoV-19/England/MILK-A05E61/2020\|EPI_ISL_600119\|20 | ···················· | ·A·················· | ···················· | ···················· | ···················· | ···················· |
| N | 7255 | hCoV-19/England/MILK-A05EF8/2020\|EPI_ISL_600120\|20 | ···················· | ···················· | ···················· | ···················· | ···················· | ···················· |
| N | 7256 | hCoV-19/England/MILK-A057A2/2020\|EPI_ISL_600121\|20 | ···················· | ·A·················· | ···················· | ···················· | ···················· | ···A················ |
| N | 7257 | hCoV-19/England/MILK-A05CD0/2020\|EPI_ISL_600122\|20 | ···················· | ···················· | ···················· | ···················· | ···················· | ···················· |
| N | 7258 | hCoV-19/England/MILK-A06334/2020\|EPI_ISL_600124\|20 | ···················· | ···················· | ···················· | ···················· | ···················· | ···················· |
| N | 7259 | hCoV-19/England/MILK-A06A7B/2020\|EPI_ISL_600125\|20 | ···················· | ···················· | ···················· | ···················· | ···················· | ···················· |
| N | 7260 | hCoV-19/Wales/MILK-A0657A/2020\|EPI_ISL_600126\|2020 | ···················· | ···················· | ···················· | ···················· | ···················· | ···················· |
| N | 7261 | hCoV-19/England/MILK-A05A5E/2020\|EPI_ISL_600127\|20 | ···················· | ···················· | ···················· | ···················· | ···················· | ···················· |
| N | 7262 | hCoV-19/England/QEUH-A00613/2020\|EPI_ISL_600128\|20 | ···················· | ···················· | ···················· | ···················· | ···················· | ···················· |
| N | 7263 | hCoV-19/Scotland/QEUH-9FFE10/2020\|EPI_ISL_600129\|2 | ···················· | ···················· | ···················· | ···················· | ···················· | ···················· |
| N | 7264 | hCoV-19/England/QEUH-9FF8A9/2020\|EPI_ISL_600130\|20 | ···················· | ···················· | ···················· | ···················· | ···················· | ···················· |
| N | 7265 | hCoV-19/England/QEUH-A00886/2020\|EPI_ISL_600131\|20 | ···················· | ···················· | ···················· | ···················· | ···················· | ···················· |
| N | 7266 | hCoV-19/Scotland/QEUH-9FF803/2020\|EPI_ISL_600132\|2 | ···················· | ···················· | ···················· | ···················· | ···················· | ···················· |
| N | 7267 | hCoV-19/England/QEUH-A00895/2020\|EPI_ISL_600133\|20 | ···················· | ···················· | ···················· | ···················· | ···················· | ···················· |
| N | 7268 | hCoV-19/England/QEUH-9FFC61/2020\|EPI_ISL_600134\|20 | ···················· | ···················· | ···················· | ···················· | ···················· | ···················· |
| N | 7269 | hCoV-19/England/QEUH-9FF3E4/2020\|EPI_ISL_600135\|20 | ···················· | ···················· | ···················· | ···················· | ···················· | ···················· |
| N | 7270 | hCoV-19/England/QEUH-A00983/2020\|EPI_ISL_600136\|20 | ···················· | ···················· | ···················· | ···················· | ···················· | ···················· |
| N | 7271 | hCoV-19/England/QEUH-A0008E/2020\|EPI_ISL_600137\|20 | ···················· | ·A·················· | ···················· | ···················· | ···················· | ···················· |
| N | 7272 | hCoV-19/England/QEUH-9FF548/2020\|EPI_ISL_600138\|20 | ···················· | ···················· | ···················· | ···················· | ···················· | ···················· |
| N | 7273 | hCoV-19/England/QEUH-9FF1AE/2020\|EPI_ISL_600140\|20 | ···················· | ···················· | ···················· | ···················· | ···················· | ···················· |
| N | 7274 | hCoV-19/Scotland/QEUH-A007A7/2020\|EPI_ISL_600141\|2 | ···················· | ···················· | ···················· | ···················· | ···················· | ···················· |
| N | 7275 | hCoV-19/Scotland/QEUH-9FFCDA/2020\|EPI_ISL_600142\|2 | ···················· | ···················· | ···················· | ···················· | ···················· | ···················· |
| N | 7276 | hCoV-19/England/QEUH-9FFD04/2020\|EPI_ISL_600143\|20 | ···················· | ···················· | ···················· | ···················· | ···················· | ···················· |
| N | 7277 | hCoV-19/England/QEUH-A0065F/2020\|EPI_ISL_600144\|20 | ···················· | ···················· | ···················· | ···················· | ······T············· | ···················· |
| N | 7278 | hCoV-19/England/QEUH-9FF593/2020\|EPI_ISL_600145\|20 | ···················· | ···················· | ···················· | ···················· | ···················· | ···················· |
| N | 7279 | hCoV-19/England/QEUH-A0033A/2020\|EPI_ISL_600146\|20 | ···················· | ···················· | ···················· | ···················· | ···················· | ···················· |
| N | 7280 | hCoV-19/Scotland/QEUH-A006C8/2020\|EPI_ISL_600147\|2 | ···················· | ···················· | ···················· | ···················· | ···················· | ···················· |
| N | 7281 | hCoV-19/England/QEUH-9FFA49/2020\|EPI_ISL_600148\|20 | ···················· | ···················· | ···················· | ···················· | ···················· | ···················· |
| N | 7282 | hCoV-19/England/QEUH-A0007F/2020\|EPI_ISL_600149\|20 | ···················· | ···················· | ···················· | ···················· | ···················· | ···················· |
| N | 7283 | hCoV-19/England/QEUH-A00200/2020\|EPI_ISL_600150\|20 | ···················· | ···················· | ···················· | ···················· | ···················· | ···················· |
| N | 7284 | hCoV-19/England/QEUH-9FFA94/2020\|EPI_ISL_600151\|20 | ···················· | ···················· | ···················· | ···················· | ···················· | ···················· |
| N | 7285 | hCoV-19/Scotland/QEUH-A006D7/2020\|EPI_ISL_600152\|2 | ···················· | ···················· | ···················· | ···················· | ···················· | ···················· |
| N | 7286 | hCoV-19/England/QEUH-9FF9E2/2020\|EPI_ISL_600153\|20 | ···················· | ···················· | ···················· | ···················· | ···················· | ···················· |
| N | 7287 | hCoV-19/England/QEUH-9FFD31/2020\|EPI_ISL_600154\|20 | ···················· | ···················· | ···················· | ···················· | ···················· | ···················· |
| N | 7288 | hCoV-19/Scotland/QEUH-A00798/2020\|EPI_ISL_600155\|2 | ···················· | ···················· | ···················· | ···················· | ···················· | ···················· |
| N | 7289 | hCoV-19/England/QEUH-A0084A/2020\|EPI_ISL_600156\|20 | ···················· | ···················· | ···················· | ···················· | ···················· | ···················· |
| N | 7290 | hCoV-19/England/QEUH-9FF3B7/2020\|EPI_ISL_600157\|20 | ···················· | ···················· | ···················· | ···················· | ···················· | ···················· |
| N | 7291 | hCoV-19/England/QEUH-9FF94C/2020\|EPI_ISL_600158\|20 | ···················· | ···················· | ···················· | ···················· | ···················· | ···················· |
| N | 7292 | hCoV-19/England/QEUH-9FFA58/2020\|EPI_ISL_600159\|20 | ···················· | ···················· | ···················· | ···················· | ···················· | ···················· |
| N | 7293 | hCoV-19/Scotland/QEUH-A007C5/2020\|EPI_ISL_600190\|2 | ···················· | ···················· | ···················· | ···················· | ···················· | ···················· |
| N | 7294 | hCoV-19/England/QEUH-A0080E/2020\|EPI_ISL_600194\|20 | ···················· | ···················· | ···················· | ···················· | ···················· | ···················· |
| N | 7295 | hCoV-19/England/QEUH-9FFCCB/2020\|EPI_ISL_600195\|20 | ···················· | ···················· | ···················· | ···················· | ···················· | ···················· |
| N | 7296 | hCoV-19/England/QEUH-9FF96A/2020\|EPI_ISL_600196\|20 | ···················· | ···················· | ···················· | ···················· | ···················· | ···················· |
| N | 7297 | hCoV-19/England/QEUH-A00297/2020\|EPI_ISL_600197\|20 | ···················· | ···················· | ···················· | ···················· | ···················· | ···················· |
| N | 7298 | hCoV-19/England/QEUH-9DA8FF/2020\|EPI_ISL_601405\|20 | ···················· | ···················· | ···················· | ···················· | ···················· | ···················· |
| N | 7299 | hCoV-19/England/MILK-9E0683/2020\|EPI_ISL_601406\|20 | ···················· | ···················· | ···················· | ···················· | ···················· | ···················· |
| N | 7300 | hCoV-19/England/QEUH-9DA7F2/2020\|EPI_ISL_601407\|20 | ···················· | ···················· | ···················· | ···················· | ···················· | ···················· |
| N | 7301 | hCoV-19/England/QEUH-9DAF36/2020\|EPI_ISL_601408\|20 | ···················· | ···················· | ···················· | ···················· | ···················· | ···················· |
| N | 7302 | hCoV-19/England/MILK-9E04E3/2020\|EPI_ISL_601409\|20 | ···················· | ···················· | ···················· | ···················· | ···················· | ···················· |
| N | 7303 | hCoV-19/England/MILK-9E05D1/2020\|EPI_ISL_601410\|20 | ···················· | ···················· | ···················· | ···················· | ···················· | ···················· |
| N | 7304 | hCoV-19/England/QEUH-9DC12F/2020\|EPI_ISL_601411\|20 | ···················· | ···················· | ···················· | ···················· | ···················· | ···················· |
| N | 7305 | hCoV-19/England/MILK-9E047A/2020\|EPI_ISL_601412\|20 | ···················· | ···················· | ···················· | ···················· | ···················· | ···················· |
| N | 7306 | hCoV-19/England/QEUH-9DA66E/2020\|EPI_ISL_601413\|20 | ···················· | ···················· | ···················· | ···················· | ···················· | ···················· |
| N | 7307 | hCoV-19/England/QEUH-9DA789/2020\|EPI_ISL_601414\|20 | ···················· | ···················· | ···················· | ···················· | ······T············· | ···················· |
| N | 7308 | hCoV-19/Wales/MILK-9E05E0/2020\|EPI_ISL_601415\|2020 | ···················· | ····A··············· | ···················· | ···················· | ···················· | ···················· |
| N | 7309 | hCoV-19/England/QEUH-9DA543/2020\|EPI_ISL_601416\|20 | ···················· | ·A·················· | ···················· | ···················· | ···················· | ···A················ |
| N | 7310 | hCoV-19/England/MILK-9E07EA/2020\|EPI_ISL_601417\|20 | ···················· | ···················· | ···················· | ···················· | ···················· | ···················· |
| N | 7311 | hCoV-19/England/MILK-9E0AE1/2020\|EPI_ISL_601418\|20 | ···················· | ···················· | ···················· | ···················· | ···················· | ···················· |
| N | 7312 | hCoV-19/Wales/MILK-9E08F6/2020\|EPI_ISL_601419\|2020 | ···················· | ···················· | ···················· | ···················· | ···················· | ···················· |
| N | 7313 | hCoV-19/England/MILK-9E0270/2020\|EPI_ISL_601420\|20 | ···················· | ···················· | ···················· | ···················· | ···················· | ···················· |
| N | 7314 | hCoV-19/England/QEUH-9DA59E/2020\|EPI_ISL_601421\|20 | ···················· | ···················· | ···················· | ···················· | ···················· | ···················· |
| N | 7315 | hCoV-19/England/QEUH-9DBE38/2020\|EPI_ISL_601423\|20 | ···················· | ···················· | ···················· | ···················· | ···················· | ···················· |
| N | 7316 | hCoV-19/England/QEUH-9DA561/2020\|EPI_ISL_601424\|20 | ···················· | ···················· | ···················· | ···················· | ···················· | ···················· |
| N | 7317 | hCoV-19/England/QEUH-9DAAAE/2020\|EPI_ISL_601426\|20 | ···················· | ···················· | ···················· | ···················· | ······T············· | ···················· |
| N | 7318 | hCoV-19/England/MILK-9E06ED/2020\|EPI_ISL_601427\|20 | ···················· | ·A·······A·········· | ···················· | ···················· | ···················· | ···················· |
| N | 7319 | hCoV-19/Wales/MILK-9E09E4/2020\|EPI_ISL_601428\|2020 | ···················· | ···················· | ···················· | ···················· | ···················· | ···················· |
| N | 7320 | hCoV-19/England/QEUH-9DBE0B/2020\|EPI_ISL_601429\|20 | ···················· | ···················· | ···················· | ···················· | ···················· | ···················· |
| N | 7321 | hCoV-19/England/QEUH-9DB06F/2020\|EPI_ISL_601430\|20 | ···················· | ···················· | ···················· | ···················· | ···················· | ···················· |
| N | 7322 | hCoV-19/England/MILK-9E06A1/2020\|EPI_ISL_601431\|20 | ···················· | ···················· | ···················· | ···················· | ···················· | ···················· |
| N | 7323 | hCoV-19/England/QEUH-9DA77A/2020\|EPI_ISL_601432\|20 | ···················· | ···················· | ···················· | ···················· | ······T············· | ···················· |
| N | 7324 | hCoV-19/England/QEUH-9DBDB3/2020\|EPI_ISL_601433\|20 | ···················· | ···················· | ···················· | ···················· | ···················· | ···················· |
| N | 7325 | hCoV-19/England/QEUH-9DB050/2020\|EPI_ISL_601436\|20 | ···················· | ···················· | ···················· | ···················· | ···················· | ···················· |

| | | | | | | | | |
|---|---|---|---|---|---|---|---|---|
| N | 7326 | hCoV-19/England/MILK-9E0BFD/2020\|EPI_ISL_601437\|20 | ···················· | ···················· | ···················· | ···················· | ···················· | ···················· |
| N | 7327 | hCoV-19/England/MILK-9E097B/2020\|EPI_ISL_601449\|20 | ···················· | ···················· | ···················· | ···················· | ···················· | ···················· |
| N | 7328 | hCoV-19/England/MILK-9E04D4/2020\|EPI_ISL_601450\|20 | ···················· | ···················· | ···················· | ···················· | ···················· | ···················· |
| N | 7329 | hCoV-19/England/MILK-9E0D33/2020\|EPI_ISL_601451\|20 | ···················· | ···················· | ···················· | ···················· | ···················· | ···················· |
| N | 7330 | hCoV-19/England/MILK-9E0498/2020\|EPI_ISL_601452\|20 | ···················· | ···················· | ···················· | ···················· | ···················· | ···················· |
| N | 7331 | hCoV-19/England/QEUH-9DA81D/2020\|EPI_ISL_601453\|20 | ···················· | ···················· | ···················· | ···················· | ····T··············· | ···················· |
| N | 7332 | hCoV-19/England/QEUH-9DA507/2020\|EPI_ISL_601454\|20 | ···················· | ·A·················· | ···················· | ···················· | ···················· | ···A················ |
| N | 7333 | hCoV-19/England/MILK-9E0D15/2020\|EPI_ISL_601455\|20 | ···················· | ···················· | ···················· | ···················· | ···················· | ···················· |
| N | 7334 | hCoV-19/England/MILK-9E0999/2020\|EPI_ISL_601456\|20 | ···················· | ···················· | ···················· | ···················· | ···················· | ···················· |
| N | 7335 | hCoV-19/Wales/MILK-9E037D/2020\|EPI_ISL_601457\|2020 | ···················· | ···················· | ···················· | ···················· | ···················· | ···················· |
| N | 7336 | hCoV-19/Wales/MILK-9E0234/2020\|EPI_ISL_601458\|2020 | ···················· | ···················· | ···················· | ···················· | ···················· | ···················· |
| N | 7337 | hCoV-19/England/QEUH-9DBF17/2020\|EPI_ISL_601459\|20 | ···················· | ···················· | ···················· | ···················· | ···················· | ···················· |
| N | 7338 | hCoV-19/England/QEUH-9DA6D7/2020\|EPI_ISL_601460\|20 | ···················· | ···················· | ···················· | ···················· | ···················· | ···················· |
| N | 7339 | hCoV-19/England/QEUH-9DBFAE/2020\|EPI_ISL_601461\|20 | ···················· | ···················· | ···················· | ···················· | ···················· | ···················· |
| N | 7340 | hCoV-19/England/QEUH-9DBDD1/2020\|EPI_ISL_601462\|20 | ···················· | ···················· | ···················· | ···················· | ···················· | ···················· |
| N | 7341 | hCoV-19/England/MILK-9E061A/2020\|EPI_ISL_601463\|20 | ···················· | ···················· | ···················· | ···················· | ···················· | ···················· |
| N | 7342 | hCoV-19/England/MILK-9E0BDF/2020\|EPI_ISL_601464\|20 | ···················· | ···················· | ···················· | ···················· | ···················· | ···················· |
| N | 7343 | hCoV-19/England/MILK-9E0A5A/2020\|EPI_ISL_601465\|20 | ···················· | ···················· | ···················· | ···················· | ···················· | ···················· |
| N | 7344 | hCoV-19/England/QEUH-9DA798/2020\|EPI_ISL_601466\|20 | ···················· | ·A·················· | ···················· | ···················· | ···················· | ···················· |
| N | 7345 | hCoV-19/England/QEUH-9DBDA4/2020\|EPI_ISL_601467\|20 | ···················· | ···················· | ···················· | ···················· | ···················· | ···················· |
| N | 7346 | hCoV-19/England/MILK-9E01BF/2020\|EPI_ISL_601468\|20 | ···················· | ···················· | ···················· | ···················· | ···················· | ···················· |
| N | 7347 | hCoV-19/England/QEUH-9DBDFF/2020\|EPI_ISL_601469\|20 | ···················· | ·A·················· | ···T················ | ···················· | ···················· | ···················· |
| N | 7348 | hCoV-19/England/MILK-9E04A7/2020\|EPI_ISL_601470\|20 | ···················· | ···················· | ···················· | ···················· | ···················· | ···················· |
| N | 7349 | hCoV-19/England/QEUH-9DBE29/2020\|EPI_ISL_601471\|20 | ···················· | ···················· | ···················· | ···················· | ···················· | ···················· |
| N | 7350 | hCoV-19/Wales/MILK-9E0B1B/2020\|EPI_ISL_601472\|2020 | ···················· | ···················· | ···················· | ···················· | ···················· | ···················· |
| N | 7351 | hCoV-19/England/MILK-9E0B57/2020\|EPI_ISL_601473\|20 | ···················· | ···················· | ···················· | ···················· | ···················· | ···················· |
| N | 7352 | hCoV-19/England/MILK-9E0331/2020\|EPI_ISL_601474\|20 | ···················· | ···················· | ···················· | ···················· | ···················· | ···················· |
| N | 7353 | hCoV-19/Wales/MILK-9E087E/2020\|EPI_ISL_601475\|2020 | ···················· | ···················· | ···················· | ···················· | ···················· | ···················· |
| N | 7354 | hCoV-19/England/MILK-9E03E6/2020\|EPI_ISL_601476\|20 | ···················· | ···················· | ···················· | ···················· | ····T··············· | ···················· |
| N | 7355 | hCoV-19/England/QEUH-9DA5BC/2020\|EPI_ISL_601477\|20 | ···················· | ···················· | ···················· | ···················· | ···················· | ···················· |
| N | 7356 | hCoV-19/England/MILK-9E03AA/2020\|EPI_ISL_601478\|20 | ···················· | ···················· | ···················· | ···················· | ···················· | ···················· |
| N | 7357 | hCoV-19/England/QEUH-9DA8C2/2020\|EPI_ISL_601479\|20 | ···················· | ···················· | ···················· | ···················· | ···················· | ···················· |
| N | 7358 | hCoV-19/England/QEUH-9DA622/2020\|EPI_ISL_601481\|20 | ···················· | ···················· | ···················· | ···················· | ···················· | ···················· |
| N | 7359 | hCoV-19/England/QEUH-9DA516/2020\|EPI_ISL_601482\|20 | ···················· | ···················· | ···················· | ···················· | ···················· | ···················· |
| N | 7360 | hCoV-19/England/QEUH-9DBD59/2020\|EPI_ISL_601483\|20 | ···················· | ···················· | ···················· | ···················· | ···················· | ···················· |
| N | 7361 | hCoV-19/Wales/MILK-9E08C9/2020\|EPI_ISL_601485\|2020 | ···················· | ···················· | ···················· | ···················· | ···················· | ···················· |
| N | 7362 | hCoV-19/England/QEUH-9DAF54/2020\|EPI_ISL_601486\|20 | ···················· | ···················· | ···················· | ···················· | ···················· | ···················· |
| N | 7363 | hCoV-19/Wales/MILK-9E0744/2020\|EPI_ISL_601487\|2020 | ···················· | ···················· | ···················· | ···················· | ···················· | ···················· |
| N | 7364 | hCoV-19/England/QEUH-9DC2C2/2020\|EPI_ISL_601488\|20 | ···················· | ···················· | ···················· | ···················· | ···················· | ···················· |
| N | 7365 | hCoV-19/England/QEUH-9DA710/2020\|EPI_ISL_601489\|20 | ···················· | ···················· | ···················· | ···················· | ···················· | ···················· |
| N | 7366 | hCoV-19/South Africa/KRISP-K003344/2020\|EPI_ISL_60 | ···················· | ·A·················· | ···················· | ···················· | ···················· | ···················· |
| N | 7367 | hCoV-19/South Africa/KRISP-K003345/2020\|EPI_ISL_60 | ···················· | ···················· | ···················· | ···················· | ···················· | ···················· |
| N | 7368 | hCoV-19/South Africa/KRISP-K003346/2020\|EPI_ISL_60 | ···················· | ···················· | ···················· | ···················· | ···················· | ···················· |
| N | 7369 | hCoV-19/South Africa/KRISP-K003347/2020\|EPI_ISL_60 | ···················· | ···················· | ···················· | ···················· | ···················· | ···················· |
| N | 7370 | hCoV-19/South Africa/KRISP-K003349/2020\|EPI_ISL_60 | ···················· | ···················· | ···················· | ···················· | ···················· | ···················· |
| N | 7371 | hCoV-19/South Africa/KRISP-K003350/2020\|EPI_ISL_60 | ···················· | ···················· | ···················· | ···················· | ···················· | ···················· |
| N | 7372 | hCoV-19/South Africa/KRISP-K003351/2020\|EPI_ISL_60 | ···················· | ···················· | ···················· | ···················· | ···················· | ···················· |
| N | 7373 | hCoV-19/South Africa/KRISP-K003352/2020\|EPI_ISL_60 | ···················· | ···················· | ···················· | ···················· | ···················· | ···················· |
| N | 7374 | hCoV-19/South Africa/KRISP-K003353/2020\|EPI_ISL_60 | ···················· | ···················· | ···················· | ···················· | ···················· | ···················· |
| N | 7375 | hCoV-19/South Africa/KRISP-K003354/2020\|EPI_ISL_60 | ···················· | ···················· | ···················· | ···················· | ···················· | ···················· |
| N | 7377 | hCoV-19/South Africa/KRISP-K003356/2020\|EPI_ISL_60 | ···················· | ···················· | ···················· | ···················· | ···················· | ···················· |
| N | 7378 | hCoV-19/South Africa/KRISP-K003357/2020\|EPI_ISL_60 | ···················· | ···················· | ···················· | ···················· | ···················· | ···················· |
| N | 7379 | hCoV-19/South Africa/KRISP-K003358/2020\|EPI_ISL_60 | ···················· | ···················· | ···················· | ···················· | ···················· | ···················· |
| N | 7380 | hCoV-19/South Africa/KRISP-K003359/2020\|EPI_ISL_60 | ···················· | ···················· | ···················· | ···················· | ···················· | ···················· |
| N | 7381 | hCoV-19/South Africa/KRISP-K003360/2020\|EPI_ISL_60 | ···················· | ···················· | ···················· | ···················· | ···················· | ···················· |
| N | 7382 | hCoV-19/South Africa/KRISP-K003361/2020\|EPI_ISL_60 | ···················· | ···················· | ···················· | ···················· | ···················· | ···················· |
| N | 7383 | hCoV-19/South Africa/KRISP-K003362/2020\|EPI_ISL_60 | ···················· | ···················· | ···················· | ···················· | ···················· | ···················· |
| N | 7384 | hCoV-19/South Africa/KRISP-K003363/2020\|EPI_ISL_60 | ···················· | ···················· | ···················· | ···················· | ···················· | ···················· |
| N | 7385 | hCoV-19/South Africa/KRISP-K003364/2020\|EPI_ISL_60 | ········T··········· | ···················· | ···················· | ···················· | ···················· | ···················· |
| N | 7386 | hCoV-19/South Africa/KRISP-K003365/2020\|EPI_ISL_60 | ···················· | ···················· | ···················· | ···················· | ···················· | ···················· |
| N | 7387 | hCoV-19/South Africa/KRISP-K003366/2020\|EPI_ISL_60 | ········T··········· | ···················· | ···················· | ···················· | ···················· | ···················· |
| N | 7388 | hCoV-19/South Africa/KRISP-K003367/2020\|EPI_ISL_60 | ········T··········· | ···················· | ···················· | ···················· | ···················· | ···················· |
| N | 7389 | hCoV-19/South Africa/KRISP-K003369/2020\|EPI_ISL_60 | ···················· | ·A·················· | ···················· | ···················· | ···················· | ···················· |
| N | 7390 | hCoV-19/South Africa/KRISP-K003371/2020\|EPI_ISL_60 | ···················· | ···················· | ···················· | ···················· | ···················· | ···················· |
| N | 7391 | hCoV-19/South Africa/KRISP-K003372/2020\|EPI_ISL_60 | ···················· | ···················· | ···················· | ···················· | ···················· | ···················· |
| N | 7392 | hCoV-19/South Africa/KRISP-K003374/2020\|EPI_ISL_60 | ···················· | ···················· | ···················· | ···················· | ···················· | ···················· |
| N | 7394 | hCoV-19/South Africa/KRISP-K003377/2020\|EPI_ISL_60 | ···················· | ···················· | ···················· | ···················· | ···················· | ···················· |
| N | 7395 | hCoV-19/South Africa/KRISP-K003378/2020\|EPI_ISL_60 | ···················· | ···················· | ···················· | ···················· | ···················· | ···················· |
| N | 7396 | hCoV-19/South Africa/KRISP-K003380/2020\|EPI_ISL_60 | ···················· | ···················· | ···················· | ···················· | ···················· | ···················· |
| N | 7397 | hCoV-19/South Africa/KRISP-K003392/2020\|EPI_ISL_60 | ···················· | ···················· | ···················· | ···················· | ···················· | ···················· |
| N | 7399 | hCoV-19/South Africa/KRISP-K003394/2020\|EPI_ISL_60 | ···················· | ···················· | ···················· | ···················· | ···················· | ···················· |
| N | 7400 | hCoV-19/South Africa/KRISP-K003395/2020\|EPI_ISL_60 | ···················· | ···················· | ···················· | ···················· | ···················· | ···················· |
| N | 7401 | hCoV-19/South Africa/KRISP-K003396/2020\|EPI_ISL_60 | ···················· | ···················· | ···················· | ···················· | ···················· | ···················· |
| N | 7402 | hCoV-19/South Africa/KRISP-K003397/2020\|EPI_ISL_60 | ···················· | ···················· | ···················· | ···················· | ···················· | ···················· |
| N | 7403 | hCoV-19/South Africa/KRISP-K003398/2020\|EPI_ISL_60 | ···················· | ···················· | ···················· | ···················· | ···················· | ···················· |
| N | 7404 | hCoV-19/South Africa/KRISP-K003399/2020\|EPI_ISL_60 | ···················· | ···················· | ···················· | ···················· | ···················· | ···················· |
| N | 7405 | hCoV-19/South Africa/KRISP-K003400/2020\|EPI_ISL_60 | ···················· | ···················· | ···················· | ···················· | ···················· | ···················· |
| N | 7406 | hCoV-19/South Africa/KRISP-K003401/2020\|EPI_ISL_60 | ········T··········· | ···················· | ···················· | ···················· | ···················· | ···················· |
| N | 7407 | hCoV-19/South Africa/KRISP-K003402/2020\|EPI_ISL_60 | ···················· | ···················· | ···················· | ···················· | ···················· | ···················· |
| N | 7408 | hCoV-19/South Africa/KRISP-K003403/2020\|EPI_ISL_60 | ···················· | ···················· | ···················· | ···················· | ···················· | ···················· |

| | | | | | | | | |
|---|---|---|---|---|---|---|---|---|
| N | 7409 | hCoV-19/South Africa/KRISP-K003459/2020\|EPI_ISL_60 | ···················· | ···················· | ···················· | ···················· | ···················· | ···················· |
| N | 7410 | hCoV-19/South Africa/KRISP-K003461/2020\|EPI_ISL_60 | ···················· | ···················· | ···················· | ···················· | ···················· | ···················· |
| N | 7411 | hCoV-19/South Africa/KRISP-K003462/2020\|EPI_ISL_60 | ···················· | ···················· | ···················· | ···················· | ···················· | ···················· |
| N | 7412 | hCoV-19/South Africa/KRISP-K003463/2020\|EPI_ISL_60 | ···················· | ·A·················· | ···················· | ···················· | ···················· | ···················· |
| N | 7413 | hCoV-19/South Africa/KRISP-K003464/2020\|EPI_ISL_60 | ···················· | ···················· | ···················· | ···················· | ···················· | ···················· |
| N | 7414 | hCoV-19/South Africa/KRISP-K003465/2020\|EPI_ISL_60 | ···················· | ···················· | ···················· | ···················· | ···················· | ···················· |
| N | 7415 | hCoV-19/USA/CA-QDX-2694/2020\|EPI_ISL_605000\|2020-0 | ···················· | ···················· | ···················· | ···················· | ···················· | ···················· |
| N | 7416 | hCoV-19/USA/CA-QDX-2697/2020\|EPI_ISL_605001\|2020-0 | ···················· | ···················· | ···················· | ···················· | ···················· | ···················· |
| N | 7417 | hCoV-19/USA/CA-QDX-2699/2020\|EPI_ISL_605002\|2020-0 | ···················· | ···················· | ···················· | ···················· | ···················· | ···················· |
| N | 7418 | hCoV-19/USA/CA-QDX-2700/2020\|EPI_ISL_605003\|2020-0 | ···················· | ···················· | ···················· | ···················· | ···················· | ···················· |
| N | 7419 | hCoV-19/USA/CA-QDX-2703/2020\|EPI_ISL_605004\|2020-0 | ···················· | ···················· | ···················· | ···················· | ···················· | ···················· |
| N | 7420 | hCoV-19/USA/CA-QDX-2705/2020\|EPI_ISL_605005\|2020-0 | ···················· | ···················· | ···················· | ···················· | ···················· | ···················· |
| N | 7421 | hCoV-19/USA/CA-QDX-2709/2020\|EPI_ISL_605006\|2020-0 | ···················· | ···················· | ···················· | ···················· | ···················· | ···················· |
| N | 7422 | hCoV-19/USA/CA-QDX-2710/2020\|EPI_ISL_605007\|2020-0 | ···················· | ···················· | ···················· | ···················· | ···················· | ···················· |
| N | 7423 | hCoV-19/USA/CA-QDX-2713/2020\|EPI_ISL_605008\|2020-0 | ···················· | ···················· | ···················· | ···················· | ···················· | ···················· |
| N | 7424 | hCoV-19/USA/CA-QDX-2718/2020\|EPI_ISL_605009\|2020-0 | ···················· | ···················· | ···················· | ···················· | ···················· | ···················· |
| N | 7425 | hCoV-19/USA/WA-S2891/2020\|EPI_ISL_596422\|2020-10-0 | ···················· | ·A·················· | ···················· | ···················· | ···················· | ···················· |
| N | 7426 | hCoV-19/USA/WA-S2892/2020\|EPI_ISL_596423\|2020-10-0 | ···················· | ···················· | ···················· | ···················· | ···················· | ···················· |
| N | 7427 | hCoV-19/USA/WA-S2895/2020\|EPI_ISL_596426\|2020-10-0 | ···················· | ···················· | ···················· | ···················· | ···················· | ···················· |
| N | 7428 | hCoV-19/USA/WA-S2896/2020\|EPI_ISL_596427\|2020-10-0 | ···················· | ···················· | ···················· | ···················· | ···················· | ···················· |
| N | 7429 | hCoV-19/USA/WA-S2899/2020\|EPI_ISL_596430\|2020-10-0 | ···················· | ·N·················· | ···················· | ···················· | ···················· | ···················· |
| N | 7430 | hCoV-19/USA/WA-S2900/2020\|EPI_ISL_596431\|2020-10-0 | ···················· | ···················· | ···················· | ···················· | ···················· | ···················· |
| N | 7431 | hCoV-19/USA/WA-S2904/2020\|EPI_ISL_596435\|2020-10-0 | ···················· | ···················· | ···················· | ···················· | ···················· | ···················· |
| N | 7432 | hCoV-19/USA/WA-S2905/2020\|EPI_ISL_596436\|2020-10-0 | ···················· | ···················· | ···················· | ···················· | ···················· | ···················· |
| N | 7433 | hCoV-19/USA/WA-S2909/2020\|EPI_ISL_596440\|2020-10-0 | ···················· | ···················· | ···················· | ···················· | ···················· | ···················· |
| N | 7434 | hCoV-19/USA/WA-S2911/2020\|EPI_ISL_596442\|2020-10-0 | ···················· | ···················· | ···················· | ···················· | ···················· | ···················· |
| N | 7435 | hCoV-19/USA/WA-S2913/2020\|EPI_ISL_596444\|2020-10-0 | ···················· | ···················· | ···················· | ···················· | ···················· | ···················· |
| N | 7436 | hCoV-19/USA/WA-S2914/2020\|EPI_ISL_596445\|2020-10-0 | ···················· | ···················· | ···················· | ···················· | ···················· | ···················· |
| N | 7437 | hCoV-19/USA/WA-S2915/2020\|EPI_ISL_596446\|2020-10-0 | ···················· | ···················· | ···················· | ···················· | ···················· | ···················· |
| N | 7438 | hCoV-19/USA/NM-LANL-2020016073/2020\|EPI_ISL_596447 | ···················· | ···················· | ···················· | ···················· | ···················· | ···················· |
| N | 7439 | hCoV-19/USA/NM-LANL-2020016074/2020\|EPI_ISL_596448 | ···················· | ···················· | ···················· | ···················· | ···················· | ···················· |
| N | 7440 | hCoV-19/Malaysia/IMR-WI085/2020\|EPI_ISL_596449\|202 | ···················· | ···················· | ···················· | ···················· | ···················· | ···················· |
| N | 7441 | hCoV-19/Iran/GRC-10582/2020\|EPI_ISL_596451\|2020-06 | ···················· | ···················· | ···················· | ···················· | ···················· | ···················· |
| N | 7442 | hCoV-19/Iran/GRC-43/2020\|EPI_ISL_596453\|2020-03-02 | ···················· | ···················· | ···················· | ···················· | ···················· | ···················· |
| N | 7443 | hCoV-19/Iran/GRC-70/2020\|EPI_ISL_596454\|2020-03-08 | ···················· | ···················· | ···················· | ···················· | ···················· | ···················· |
| N | 7444 | hCoV-19/Iran/GRC-9695/2020\|EPI_ISL_596455\|2020-06- | ···················· | ···················· | ···················· | ···················· | ···················· | ···················· |
| N | 7445 | hCoV-19/Singapore/1066/2020\|EPI_ISL_596456\|2020-10 | ···················· | ·A·················· | ···················· | ···················· | ···················· | ···A················ |
| N | 7446 | hCoV-19/Singapore/1067/2020\|EPI_ISL_596457\|2020-10 | ···················· | ···················· | ···················· | ···················· | ···················· | ···················· |
| N | 7447 | hCoV-19/Singapore/1068/2020\|EPI_ISL_596458\|2020-10 | ···················· | ···················· | ···················· | ···················· | ···················· | ···················· |
| N | 7448 | hCoV-19/Singapore/1069/2020\|EPI_ISL_596459\|2020-10 | ···················· | ···················· | ···················· | ···················· | ···················· | ···················· |
| N | 7449 | hCoV-19/Singapore/1070/2020\|EPI_ISL_596460\|2020-10 | ···················· | ···················· | ···················· | ···················· | ···················· | ···················· |
| N | 7450 | hCoV-19/Singapore/1071/2020\|EPI_ISL_596461\|2020-10 | ···················· | ·A·················· | ···················· | ···················· | ···················· | ···A················ |
| N | 7451 | hCoV-19/Singapore/1072/2020\|EPI_ISL_596462\|2020-10 | ···················· | ···················· | ···················· | ···················· | ···················· | ···················· |
| N | 7452 | hCoV-19/Singapore/1073/2020\|EPI_ISL_596463\|2020-09 | ···················· | ···················· | ···················· | ···················· | ···················· | ···················· |
| N | 7453 | hCoV-19/Singapore/1074/2020\|EPI_ISL_596464\|2020-10 | ···················· | ·A·················· | ···················· | ···················· | ···················· | ···················· |
| N | 7454 | hCoV-19/Singapore/1075/2020\|EPI_ISL_596465\|2020-04 | ···················· | ···················· | ···················· | ···················· | ···················· | ···················· |
| N | 7455 | hCoV-19/England/ALDP-A50AAF/2020\|EPI_ISL_597710\|20 | ···················· | ···················· | ···················· | ···················· | ···················· | ···················· |
| N | 7456 | hCoV-19/England/ALDP-A502C5/2020\|EPI_ISL_597711\|20 | ···················· | ···················· | ···················· | ···················· | ···················· | ···················· |
| N | 7457 | hCoV-19/England/ALDP-A50359/2020\|EPI_ISL_597713\|20 | ···················· | ···················· | ···················· | ···················· | ···················· | ···················· |
| N | 7458 | hCoV-19/England/ALDP-A4FDB9/2020\|EPI_ISL_597714\|20 | ···················· | ···················· | ···················· | ···················· | ···················· | ···················· |
| N | 7459 | hCoV-19/England/ALDP-A5031D/2020\|EPI_ISL_597715\|20 | ···················· | ···················· | ···················· | ···················· | ···················· | ···················· |
| N | 7460 | hCoV-19/England/ALDP-A50A63/2020\|EPI_ISL_597716\|20 | ···················· | ···················· | ···················· | ···················· | ···················· | ···················· |
| N | 7461 | hCoV-19/England/ALDP-A4FD13/2020\|EPI_ISL_597720\|20 | ···················· | ···················· | ···················· | ···················· | ···················· | ···················· |
| N | 7462 | hCoV-19/England/ALDP-A50104/2020\|EPI_ISL_597721\|20 | ···················· | ·A·················· | ···················· | ···················· | ···················· | ···A················ |
| N | 7463 | hCoV-19/England/ALDP-A50517/2020\|EPI_ISL_597722\|20 | ···················· | ···················· | ···················· | ···················· | ···················· | ···················· |
| N | 7464 | hCoV-19/England/ALDP-A50492/2020\|EPI_ISL_597723\|20 | ···················· | ···················· | ···················· | ···················· | ···················· | ···················· |
| N | 7465 | hCoV-19/England/ALDP-A5026B/2020\|EPI_ISL_597724\|20 | ···················· | ···················· | ···················· | ···················· | ···················· | ···················· |
| N | 7466 | hCoV-19/England/ALDP-A5030E/2020\|EPI_ISL_597725\|20 | ···················· | ···················· | ···················· | ···················· | ···················· | ···················· |
| N | 7467 | hCoV-19/England/ALDP-A50BBB/2020\|EPI_ISL_597726\|20 | ···················· | ···················· | ···················· | ···················· | ···················· | ···················· |
| N | 7468 | hCoV-19/England/ALDP-A5078A/2020\|EPI_ISL_597727\|20 | ···················· | ···················· | ···················· | ···················· | ···················· | ···················· |
| N | 7469 | hCoV-19/England/ALDP-A50D6A/2020\|EPI_ISL_597728\|20 | ···················· | ···················· | ···················· | ···················· | ···················· | ···················· |
| N | 7470 | hCoV-19/England/ALDP-A4FB46/2020\|EPI_ISL_597729\|20 | ···················· | ···················· | ···················· | ···················· | ···················· | ···················· |
| N | 7471 | hCoV-19/England/ALDP-A50B06/2020\|EPI_ISL_597730\|20 | ···················· | ···················· | ···················· | ···················· | ···················· | ···················· |
| N | 7472 | hCoV-19/England/ALDP-A50395/2020\|EPI_ISL_597756\|20 | ···················· | ···················· | ···················· | ···················· | ···················· | ···················· |
| N | 7473 | hCoV-19/England/ALDP-A50605/2020\|EPI_ISL_597757\|20 | ···················· | ·········A·········· | ···················· | ···················· | ···················· | ···················· |
| N | 7474 | hCoV-19/England/ALDP-A50C6D/2020\|EPI_ISL_597758\|20 | ···················· | ···················· | ···················· | ···················· | ···················· | ···················· |
| N | 7475 | hCoV-19/England/ALDP-A50F28/2020\|EPI_ISL_597759\|20 | ···················· | ···················· | ···················· | ···················· | ···················· | ···················· |
| N | 7476 | hCoV-19/England/ALDP-A5069C/2020\|EPI_ISL_597760\|20 | ···················· | ···················· | ···················· | ···················· | ···················· | ···················· |
| N | 7477 | hCoV-19/England/ALDP-A4FDF5/2020\|EPI_ISL_597761\|20 | ···················· | ·A·················· | ···················· | ···················· | ···················· | ···A················ |
| N | 7478 | hCoV-19/England/ALDP-A50A27/2020\|EPI_ISL_597762\|20 | ···················· | ···················· | ···················· | ···················· | ···················· | ···················· |
| N | 7479 | hCoV-19/England/ALDP-A4FB37/2020\|EPI_ISL_597764\|20 | ···················· | ···················· | ···················· | ···················· | ···················· | ···················· |
| N | 7480 | hCoV-19/England/ALDP-A4FD40/2020\|EPI_ISL_597765\|20 | ···················· | ···················· | ···················· | ···················· | ···················· | ···················· |
| N | 7481 | hCoV-19/England/ALDP-A50FCE/2020\|EPI_ISL_597766\|20 | ···················· | ···················· | ···················· | ···················· | ···················· | ···················· |
| N | 7482 | hCoV-19/England/ALDP-A50F37/2020\|EPI_ISL_597767\|20 | ···················· | ··············C····· | ···················· | ···················· | ···················· | ···················· |
| N | 7483 | hCoV-19/England/ALDP-A4FF59/2020\|EPI_ISL_597769\|20 | ···················· | ···················· | ···················· | ···················· | ···················· | ···················· |
| N | 7484 | hCoV-19/England/ALDP-A4FDD7/2020\|EPI_ISL_597770\|20 | ···················· | ···················· | ···················· | ···················· | ···················· | ···················· |
| N | 7485 | hCoV-19/England/ALDP-A50580/2020\|EPI_ISL_597771\|20 | ···················· | ···················· | ···················· | ···················· | ···················· | ···················· |
| N | 7486 | hCoV-19/England/ALDP-A507B7/2020\|EPI_ISL_597772\|20 | ···················· | ···················· | ···················· | ···················· | ···················· | ···················· |
| N | 7487 | hCoV-19/England/ALDP-A502F2/2020\|EPI_ISL_597773\|20 | ···················· | ···················· | ···················· | ···················· | ···················· | ···················· |
| N | 7488 | hCoV-19/England/ALDP-A4FA3A/2020\|EPI_ISL_597774\|20 | ···················· | ···················· | ···················· | ···················· | ···················· | ···················· |

```
N 7489 hCoV-19/England/ALDP-A4FDE6/2020|EPI_ISL_597775|20 .................... .................... .................... .................... .................... ....................
N 7490 hCoV-19/England/ALDP-A5025C/2020|EPI_ISL_597776|20 .................... .................... .................... .................... .................... ....................
N 7491 hCoV-19/England/ALDP-A502B6/2020|EPI_ISL_597778|20 .................... .................... .................... .................... .................... ....................
N 7492 hCoV-19/England/ALDP-A4FAEF/2020|EPI_ISL_597779|20 .................... .................... .................... .................... .................... ....................
N 7493 hCoV-19/England/ALDP-A4FC16/2020|EPI_ISL_597780|20 .................... .................... .................... .................... .................... ....................
N 7494 hCoV-19/England/ALDP-A504CF/2020|EPI_ISL_597781|20 .................... .................... .................... .................... .................... ....................
N 7495 hCoV-19/England/ALDP-A50D2E/2020|EPI_ISL_597782|20 .................... .................... .................... .................... .................... ....................
N 7496 hCoV-19/England/ALDP-A50993/2020|EPI_ISL_597783|20 .................... .................... .................... .................... .................... ....................
N 7497 hCoV-19/England/ALDP-A5073F/2020|EPI_ISL_597784|20 .................... .................... .................... .................... .................... ....................
N 7498 hCoV-19/England/ALDP-A50016/2020|EPI_ISL_597785|20 .................... .................... .................... .................... .................... ....................
N 7499 hCoV-19/England/ALDP-A506C9/2020|EPI_ISL_597786|20 .................... .................... .................... .................... .................... ....................
N 7500 hCoV-19/England/ALDP-A504DE/2020|EPI_ISL_597787|20 .................... .................... .................... .................... .................... ....................
N 7501 hCoV-19/England/ALDP-A5023E/2020|EPI_ISL_597789|20 .................... .................... .................... .................... .................... ....................
N 7502 hCoV-19/England/ALDP-A50C21/2020|EPI_ISL_597790|20 .................... .................... .................... .................... .................... ....................
N 7503 hCoV-19/England/ALDP-A50A18/2020|EPI_ISL_597791|20 .................... .A.................. .................... .................... .................... ..A.................
N 7504 hCoV-19/England/ALDP-A509EE/2020|EPI_ISL_597792|20 .................... .................... .................... .................... .................... ..A.................
N 7505 hCoV-19/England/ALDP-A5114F/2020|EPI_ISL_597793|20 .................... .................... .................... .................... .................... ....................
N 7506 hCoV-19/Wales/ALDP-A4FF3B/2020|EPI_ISL_597794|2020 .................... .................... .................... .................... .................... ....................
N 7507 hCoV-19/England/ALDP-A50A90/2020|EPI_ISL_597795|20 .................... .................... .................... .................... .................... ....................
N 7508 hCoV-19/England/ALDP-A50508/2020|EPI_ISL_597796|20 .................... .................... .................... .................... .................... ....................
N 7509 hCoV-19/Scotland/QEUH-A1E06F/2020|EPI_ISL_598905|2 .................... .................... .................... .................... .................... ....................
N 7510 hCoV-19/Scotland/QEUH-A1E9EC/2020|EPI_ISL_598906|2 .................... .................... .................... .................... .................... ....................
N 7511 hCoV-19/Scotland/QEUH-A1E436/2020|EPI_ISL_598907|2 .................... .................... .................... .................... .................... ....................
N 7512 hCoV-19/Scotland/QEUH-A1DD5A/2020|EPI_ISL_598908|2 .................... .................... .................... .................... .................... ....................
N 7513 hCoV-19/Scotland/QEUH-A21861/2020|EPI_ISL_598909|2 .................... .................... .................... .................... .................... ....................
N 7514 hCoV-19/England/MILK-A068EA/2020|EPI_ISL_600000|20 .................... .................... .................... .................... .................... ....................
N 7515 hCoV-19/England/MILK-A066B3/2020|EPI_ISL_600001|20 .................... .................... .................... .................... .................... ....................
N 7516 hCoV-19/England/MILK-A053DB/2020|EPI_ISL_600002|20 .................... .A.................. .................... .................... .................... ..A.................
N 7517 hCoV-19/England/MILK-A05EDA/2020|EPI_ISL_600003|20 .................... .................... .................... .................... .................... ....................
N 7518 hCoV-19/England/MILK-A05696/2020|EPI_ISL_600005|20 .................... .................... .................... .................... .................... ....................
N 7519 hCoV-19/Wales/MILK-A063F8/2020|EPI_ISL_600006|2020 .................... .................... .................... .................... .................... ....................
N 7520 hCoV-19/England/MILK-A0645F/2020|EPI_ISL_600040|20 .................... .................... .................... .................... .................... ....................
N 7521 hCoV-19/England/MILK-A057DF/2020|EPI_ISL_600041|20 .................... .................... .................... .................... .................... ....................
N 7522 hCoV-19/England/MILK-A0601F/2020|EPI_ISL_600042|20 .................... .................... .................... .................... .................... ....................
N 7523 hCoV-19/England/MILK-A064B9/2020|EPI_ISL_600043|20 .................... .................... .................... .................... .................... ....................
N 7524 hCoV-19/England/MILK-A06361/2020|EPI_ISL_600044|20 .................... .................... .................... .................... ......T............. ....................
N 7525 hCoV-19/England/MILK-A068AE/2020|EPI_ISL_600045|20 .................... .................... .................... .................... .................... ....................
N 7526 hCoV-19/England/MILK-A0556C/2020|EPI_ISL_600046|20 ...........T........ .................... .................... .................... .................... ....................
N 7527 hCoV-19/England/MILK-A05B3D/2020|EPI_ISL_600048|20 .................... .................... .................... .................... .................... ....................
N 7528 hCoV-19/England/MILK-A054E7/2020|EPI_ISL_600049|20 .................... .................... .................... .................... ......T............. ....................
N 7529 hCoV-19/Wales/MILK-A05B4C/2020|EPI_ISL_600050|2020 .................... .................... .................... .................... .................... ....................
N 7530 hCoV-19/England/MILK-A05E16/2020|EPI_ISL_600051|20 .................... .A.................. .................... .................... .................... ....................
N 7531 hCoV-19/England/MILK-A05739/2020|EPI_ISL_600052|20 .................... .................... .................... .................... ......T............. ....................
N 7532 hCoV-19/England/MILK-A0664A/2020|EPI_ISL_600054|20 .................... .................... .................... .................... .................... ....................
N 7533 hCoV-19/England/MILK-A0670B/2020|EPI_ISL_600055|20 .................... .................... .................... .................... .................... ....................
N 7534 hCoV-19/England/MILK-A056B4/2020|EPI_ISL_600057|20 .................... .A.................. .................... .................... .................... ..A.................
N 7535 hCoV-19/England/MILK-A06088/2020|EPI_ISL_600058|20 .................... .A.................. .................... .................... .................... ..A.................
N 7536 hCoV-19/England/MILK-A053CC/2020|EPI_ISL_600059|20 .................... .................... .................... .................... .................... ....................
N 7537 hCoV-19/England/MILK-A06A5D/2020|EPI_ISL_600060|20 .................... .................... .................... .................... .................... ....................
N 7538 hCoV-19/England/MILK-A0560F/2020|EPI_ISL_600061|20 .................... .................... .................... .................... .................... ....................
N 7539 hCoV-19/England/MILK-A0648C/2020|EPI_ISL_600062|20 .................... .A.................. .................... .................... .................... ..A.................
N 7540 hCoV-19/England/MILK-A05E8F/2020|EPI_ISL_600063|20 .................... .................... .................... .................... ......T............. ....................
N 7541 hCoV-19/Wales/MILK-A05D82/2020|EPI_ISL_600064|2020 .................... .................... .................... .................... .................... ....................
N 7542 hCoV-19/England/MILK-A06273/2020|EPI_ISL_600065|20 .................... .................... .................... .................... .................... ....................
N 7543 hCoV-19/England/MILK-A0649B/2020|EPI_ISL_600066|20 .................... .R.................. .................... .................... .................... ....................
N 7544 hCoV-19/Wales/MILK-A069F6/2020|EPI_ISL_600067|2020 .................... .................... .................... .................... .................... ....................
N 7545 hCoV-19/England/MILK-A055C6/2020|EPI_ISL_600068|20 .................... .................... .................... .................... .................... ....................
N 7546 hCoV-19/England/MILK-A06255/2020|EPI_ISL_600069|20 .................... .A.................. .................... .................... .................... ....................
N 7547 hCoV-19/England/MILK-A05809/2020|EPI_ISL_600070|20 .................... .................... .................... .................... .................... ....................
N 7548 hCoV-19/England/MILK-A064D7/2020|EPI_ISL_600071|20 .................... .................... .................... .................... .................... ....................
N 7549 hCoV-19/England/MILK-A0662C/2020|EPI_ISL_600072|20 .................... .................... .................... .................... .................... ....................
N 7550 hCoV-19/England/MILK-A063BC/2020|EPI_ISL_600073|20 .................... .................... .................... .................... .................... ....................
N 7551 hCoV-19/England/MILK-A05CA3/2020|EPI_ISL_600074|20 .................... .................... .................... .................... .................... ....................
N 7552 hCoV-19/England/MILK-A05DFB/2020|EPI_ISL_600075|20 .................... .................... .................... .................... .................... ....................
N 7553 hCoV-19/England/MILK-A05D46/2020|EPI_ISL_600076|20 .................... .................... .................... .................... .................... ....................
N 7554 hCoV-19/England/MILK-A062BF/2020|EPI_ISL_600077|20 .................... .................... .................... .................... .................... ....................
N 7555 hCoV-19/England/MILK-A0558A/2020|EPI_ISL_600078|20 .................... .................... .................... .................... .................... ....................
N 7556 hCoV-19/England/MILK-A064E6/2020|EPI_ISL_600079|20 .................... .................... .................... .................... .................... ....................
N 7557 hCoV-19/England/MILK-A05C94/2020|EPI_ISL_600103|20 .................... .................... .................... .................... .................... ....................
N 7558 hCoV-19/England/MILK-A05933/2020|EPI_ISL_600104|20 .................... .A.................. .................... .................... .................... ....................
N 7559 hCoV-19/Wales/MILK-A065A7/2020|EPI_ISL_600108|2020 .................... .................... .................... .................... .................... ....................
N 7560 hCoV-19/England/MILK-A059AC/2020|EPI_ISL_600109|20 .................... .A.................. .................... .................... .................... ..A.................
N 7561 hCoV-19/England/MILK-A05BB5/2020|EPI_ISL_600110|20 .................... .................... .................... .................... .................... ....................
N 7562 hCoV-19/England/MILK-A06228/2020|EPI_ISL_600111|20 .................... .A.................. .................... .................... .................... ..A.................
N 7563 hCoV-19/England/MILK-A0570C/2020|EPI_ISL_600112|20 .................... .................... .................... .................... .................... ....................
N 7564 hCoV-19/England/MILK-A05599/2020|EPI_ISL_600113|20 .................... .................... .................... .................... .................... ....................
N 7565 hCoV-19/England/MILK-A060C4/2020|EPI_ISL_600114|20 .................... .................... .................... .................... .................... ....................
N 7566 hCoV-19/England/MILK-A06422/2020|EPI_ISL_600115|20 .................... .................... .................... .................... .................... ....................
N 7567 hCoV-19/England/MILK-A0613A/2020|EPI_ISL_600116|20 .................... .................... .................... .................... .................... ....................
N 7568 hCoV-19/England/MILK-A06219/2020|EPI_ISL_600117|20 .................... .................... .................... .................... .................... ....................
```

```
N  7569 hCoV-19/England/MILK-9E0AB4/2020|EPI_ISL_601329|20    ....................  ....................  ....................  ....................  ....................  ....................
N  7570 hCoV-19/England/QEUH-9DA74D/2020|EPI_ISL_601331|20    ....................  ....................  ....................  ....................  ....................  ....................
N  7571 hCoV-19/England/QEUH-9DA4EC/2020|EPI_ISL_601332|20    ....................  ....................  ....................  ....................  ....................  ....................
N  7572 hCoV-19/England/QEUH-9DBF08/2020|EPI_ISL_601333|20    ....................  ....................  ....................  ....................  ....................  ....................
N  7573 hCoV-19/England/QEUH-9DBE1A/2020|EPI_ISL_601334|20    ....................  .A..................  ....................  ....................  ....................  ....................
N  7574 hCoV-19/England/QEUH-9DA886/2020|EPI_ISL_601336|20    ....................  ....................  ....................  ....................  ....................  ....................
N  7575 hCoV-19/Wales/MILK-9E0805/2020|EPI_ISL_601337|2020     ....................  ....................  ....................  ....................  ....................  ....................
N  7576 hCoV-19/England/MILK-9E0AD2/2020|EPI_ISL_601338|20    ....................  ....................  ....................  ....................  ....................  ....................
N  7577 hCoV-19/England/MILK-9E043E/2020|EPI_ISL_601370|20    ....................  .A.......A..........  ....................  ....................  ....................  ....................
N  7578 hCoV-19/England/MILK-9E0CCD/2020|EPI_ISL_601371|20    ....................  ....................  ....................  ....................  ....................  ....................
N  7579 hCoV-19/England/QEUH-9DC2E0/2020|EPI_ISL_601372|20    ....................  ....................  ....................  ....................  ....................  ....................
N  7580 hCoV-19/England/MILK-9E044D/2020|EPI_ISL_601373|20    ....................  ....................  ....................  ....................  ....................  ....................
N  7581 hCoV-19/England/QEUH-9DA895/2020|EPI_ISL_601374|20    ....................  ....................  ....................  ....................  ....................  ....................
N  7582 hCoV-19/England/MILK-9E0D06/2020|EPI_ISL_601376|20    ....................  ....................  ....................  ....................  ....................  ....................
N  7583 hCoV-19/Wales/MILK-9E0C63/2020|EPI_ISL_601378|2020     ....................  ....................  ....................  ....................  ....................  ....................
N  7584 hCoV-19/England/MILK-9E09A8/2020|EPI_ISL_601379|20    ....................  ....................  ....................  ....................  ....................  ....................
N  7585 hCoV-19/England/QEUH-9DBFF9/2020|EPI_ISL_601380|20    ....................  ....................  ....................  ....................  ....................  ....................
N  7586 hCoV-19/Wales/MILK-9E0B0C/2020|EPI_ISL_601381|2020     ....................  ....................  ....................  ....................  ....................  ....................
N  7587 hCoV-19/Wales/MILK-9E0D7F/2020|EPI_ISL_601382|2020     ....................  ....................  ....................  ....................  ....................  ....................
N  7588 hCoV-19/England/MILK-9E08AB/2020|EPI_ISL_601383|20    ....................  ....................  ....................  ....................  ....................  ....................
N  7589 hCoV-19/England/QEUH-9DC2FF/2020|EPI_ISL_601384|20    ....................  ....................  ....................  ....................  ....................  ....................
N  7590 hCoV-19/Wales/MILK-9E0726/2020|EPI_ISL_601385|2020     ....................  ....................  ....................  ....................  ....................  ....................
N  7591 hCoV-19/England/QEUH-9DBD68/2020|EPI_ISL_601386|20    ....................  ....................  ....................  ....................  ....................  ....................
N  7592 hCoV-19/England/QEUH-9DC040/2020|EPI_ISL_601387|20    ....................  .A..................  ....................  ....................  ....................  ..A.................
N  7593 hCoV-19/England/MILK-9E0568/2020|EPI_ISL_601388|20    ....................  ....................  ....................  ....................  ....................  ....................
N  7594 hCoV-19/England/QEUH-9DBE47/2020|EPI_ISL_601389|20    ....................  ....................  ....................  ....................  ....................  ....................
N  7595 hCoV-19/England/MILK-9E06DE/2020|EPI_ISL_601390|20    ....................  ....................  ....................  ....................  ....................  ....................
N  7596 hCoV-19/England/QEUH-9DAF27/2020|EPI_ISL_601391|20    ....................  .A..................  ....................  ....................  ....................  ..A.................
N  7597 hCoV-19/England/MILK-9E035F/2020|EPI_ISL_601392|20    ....................  ....................  ....................  ....................  ....................  ....................
N  7598 hCoV-19/England/QEUH-9DC295/2020|EPI_ISL_601393|20    ....................  ....................  ....................  ....................  ....................  ....................
N  7599 hCoV-19/England/MILK-9E0C36/2020|EPI_ISL_601394|20    ....................  ....................  ....................  ....................  ....................  ....................
N  7600 hCoV-19/Wales/MILK-9E0814/2020|EPI_ISL_601396|2020     ....................  ....................  ....................  ....................  ....................  ....................
N  7601 hCoV-19/Wales/MILK-9E0207/2020|EPI_ISL_601397|2020     ....................  ....................  ....................  ....................  ....................  ....................
N  7602 hCoV-19/Wales/MILK-9E0C45/2020|EPI_ISL_601398|2020     ....................  ....................  ....................  ....................  ....................  ....................
N  7603 hCoV-19/England/QEUH-9DA67D/2020|EPI_ISL_601400|20    ....................  ....................  ....................  ....................  ....................  ....................
N  7604 hCoV-19/England/QEUH-9DA90B/2020|EPI_ISL_601401|20    ....................  ....................  ....................  ....................  ....................  ....................
N  7605 hCoV-19/England/QEUH-9DA965/2020|EPI_ISL_601402|20    ....................  ....................  ....................  ....................  ....................  ....................
N  7606 hCoV-19/England/QEUH-9DC013/2020|EPI_ISL_601403|20    ....................  ....................  ....................  ....................  ....................  ....................
N  7607 hCoV-19/England/MILK-9E0CAF/2020|EPI_ISL_601404|20    ....................  ....................  ....................  ....................  ....................  ....................
N  7608 hCoV-19/England/MILK-9E0BB1/2020|EPI_ISL_601434|20    ....................  ....................  ....................  ....................  ....................  ....................
N  7609 hCoV-19/England/QEUH-9DA701/2020|EPI_ISL_601435|20    ....................  ....................  ....................  ....................  ....................  ....................
N  7610 hCoV-19/England/MILK-9E0595/2020|EPI_ISL_601438|20    ....................  .A..................  ....................  ....................  ....................  ..A.................
N  7611 hCoV-19/England/MILK-9E0CBE/2020|EPI_ISL_601439|20    ....................  ....................  ....................  ....................  ....................  ....................
N  7612 hCoV-19/Wales/MILK-9E0C54/2020|EPI_ISL_601440|2020     ....................  ....................  ....................  ....................  ....................  ....................
N  7613 hCoV-19/Wales/MILK-9E0252/2020|EPI_ISL_601441|2020     ....................  ....................  ....................  ....................  ....................  ....................
N  7614 hCoV-19/England/MILK-9E0A87/2020|EPI_ISL_601442|20    ....................  ....................  ....................  ....................  ....................  ....................
N  7615 hCoV-19/England/MILK-9E0780/2020|EPI_ISL_601444|20    ....................  ....................  ....................  ....................  ....................  ....................
N  7616 hCoV-19/England/QEUH-9DA4CE/2020|EPI_ISL_601445|20    ....................  ....................  ....................  ....................  ......T.............  ....................
N  7617 hCoV-19/England/QEUH-9DA72F/2020|EPI_ISL_601446|20    ....................  ....................  ....................  ....................  ....................  ....................
N  7618 hCoV-19/England/MILK-9E04B6/2020|EPI_ISL_601447|20    ....................  ....................  ....................  ....................  ....................  ....................
N  7619 hCoV-19/England/MILK-9E02F8/2020|EPI_ISL_601448|20    ....................  ....................  ....................  ....................  ....................  ....................
N  7620 hCoV-19/South Africa/KRISP-K002843/2020|EPI_ISL_60    ....................  ....................  ....................  ....................  ....................  ....................
N  7621 hCoV-19/South Africa/KRISP-K002846/2020|EPI_ISL_60    ....................  ....................  ....................  ....................  ....................  ....................
N  7622 hCoV-19/South Africa/KRISP-K002847/2020|EPI_ISL_60    ....................  ....................  ....................  ....................  ....................  ....................
N  7623 hCoV-19/South Africa/KRISP-K002848/2020|EPI_ISL_60    ....................  ....................  ....................  ....................  ....................  ....................
N  7624 hCoV-19/South Africa/KRISP-K002849/2020|EPI_ISL_60    ....................  .A..................  ....................  ....................  ....................  ....................
N  7625 hCoV-19/South Africa/KRISP-K002850/2020|EPI_ISL_60    ....................  ....................  ....................  ....................  ....................  ....................
N  7626 hCoV-19/South Africa/KRISP-K002851/2020|EPI_ISL_60    ....................  ....................  ....................  ....................  ....................  ....................
N  7627 hCoV-19/South Africa/KRISP-K002852/2020|EPI_ISL_60    ....................  ....................  ....................  ....................  ....................  ....................
N  7628 hCoV-19/South Africa/KRISP-K002853/2020|EPI_ISL_60    ....................  ....................  ....................  ....................  ....................  ....................
N  7629 hCoV-19/South Africa/KRISP-K002888/2020|EPI_ISL_60    ....................  ....................  ....................  ....................  ....................  ....................
N  7630 hCoV-19/South Africa/KRISP-K002889/2020|EPI_ISL_60    ....................  ....................  ....................  ....................  ....................  ....................
N  7631 hCoV-19/South Africa/KRISP-K002891/2020|EPI_ISL_60    ....................  ....................  ....................  ....................  ....................  ....................
N  7632 hCoV-19/South Africa/KRISP-K002892/2020|EPI_ISL_60    ....................  ....................  ....................  ....................  ....................  ....................
N  7633 hCoV-19/South Africa/KRISP-K002893/2020|EPI_ISL_60    ....................  ....................  ....................  ....................  ....................  ....................
N  7634 hCoV-19/South Africa/KRISP-K002894/2020|EPI_ISL_60    ....................  ....................  ....................  ....................  ....................  ....................
N  7635 hCoV-19/South Africa/KRISP-K002895/2020|EPI_ISL_60    ....................  ....................  ....................  ....................  ....................  ....................
N  7636 hCoV-19/South Africa/KRISP-K002896/2020|EPI_ISL_60    ....................  ....................  ....................  ....................  ....................  ....................
N  7637 hCoV-19/South Africa/KRISP-K002897/2020|EPI_ISL_60    ....................  ....................  ....................  ....................  ....................  ....................
N  7638 hCoV-19/South Africa/KRISP-K002898/2020|EPI_ISL_60    ....................  ....................  ....................  ....................  ....................  ....................
N  7639 hCoV-19/South Africa/KRISP-K002899/2020|EPI_ISL_60    ....................  ....................  ....................  ....................  ....................  ....................
N  7640 hCoV-19/South Africa/KRISP-K002901/2020|EPI_ISL_60    ....................  ....................  ....................  ....................  ....................  ....................
N  7641 hCoV-19/South Africa/KRISP-K002902/2020|EPI_ISL_60    ....................  ....................  ....................  ....................  ....................  ....................
N  7642 hCoV-19/South Africa/KRISP-K002903/2020|EPI_ISL_60    ....................  ....................  ....................  ....................  ....................  ....................
N  7643 hCoV-19/South Africa/KRISP-K003322/2020|EPI_ISL_60    ....................  ....................  ....................  ....................  ....................  ....................
N  7644 hCoV-19/South Africa/KRISP-K003323/2020|EPI_ISL_60    ....................  ....................  ....................  ....................  ....................  ....................
N  7645 hCoV-19/South Africa/KRISP-K003324/2020|EPI_ISL_60    ....................  ....................  ....................  ....................  ....................  ....................
N  7646 hCoV-19/South Africa/KRISP-K003325/2020|EPI_ISL_60    ....................  .A..................  ....................  ....................  ....................  ....................
N  7647 hCoV-19/South Africa/KRISP-K003326/2020|EPI_ISL_60    ....................  ....................  ....................  ....................  ....................  ....................
N  7648 hCoV-19/South Africa/KRISP-K003327/2020|EPI_ISL_60    ....................  ....................  ....................  ....................  ....................  ....................
```

| | | | | | | | | |
|---|---|---|---|---|---|---|---|---|
| N | 7649 | hCoV-19/South Africa/KRISP-K003328/2020\|EPI_ISL_60 | ···················· | ···················· | ···················· | ···················· | ···················· | ···················· |
| N | 7651 | hCoV-19/South Africa/KRISP-K003330/2020\|EPI_ISL_60 | ···················· | ···················· | ···················· | ···················· | ···················· | ···················· |
| N | 7652 | hCoV-19/South Africa/KRISP-K003331/2020\|EPI_ISL_60 | ···················· | ···················· | ···················· | ···················· | ···················· | ···················· |
| N | 7653 | hCoV-19/South Africa/KRISP-K003332/2020\|EPI_ISL_60 | ···················· | ···················· | ···················· | ···················· | ···················· | ···················· |
| N | 7654 | hCoV-19/South Africa/KRISP-K003334/2020\|EPI_ISL_60 | ···················· | ···················· | ···················· | ···················· | ···················· | ···················· |
| N | 7655 | hCoV-19/South Africa/KRISP-K003335/2020\|EPI_ISL_60 | ···················· | ···················· | ···················· | ···················· | ···················· | ···················· |
| N | 7656 | hCoV-19/South Africa/KRISP-K003336/2020\|EPI_ISL_60 | ···················· | ···················· | ···················· | ···················· | ···················· | ···················· |
| N | 7657 | hCoV-19/South Africa/KRISP-K003337/2020\|EPI_ISL_60 | ···················· | ···················· | ···················· | ···················· | ···················· | ···················· |
| N | 7658 | hCoV-19/South Africa/KRISP-K003338/2020\|EPI_ISL_60 | ···················· | ···················· | ···················· | ···················· | ···················· | ···················· |
| N | 7659 | hCoV-19/South Africa/KRISP-K003339/2020\|EPI_ISL_60 | ···················· | ···················· | ···················· | ···················· | ···················· | ···················· |
| N | 7660 | hCoV-19/South Africa/KRISP-K003340/2020\|EPI_ISL_60 | ···················· | ······A············· | ···················· | ···················· | ···················· | ···················· |
| N | 7661 | hCoV-19/South Africa/KRISP-K003341/2020\|EPI_ISL_60 | ···················· | ···················· | ···················· | ···················· | ···················· | ···················· |
| N | 7662 | hCoV-19/South Africa/KRISP-K003343/2020\|EPI_ISL_60 | ···················· | ···················· | ···················· | ···················· | ···················· | ···················· |
| N | 7663 | hCoV-19/South Africa/KRISP-K003370/2020\|EPI_ISL_60 | ···················· | ···················· | ···················· | ···················· | ···················· | ···················· |
| N | 7664 | hCoV-19/South Africa/KRISP-K003375/2020\|EPI_ISL_60 | ···················· | ···················· | ···················· | ···················· | ···················· | ···················· |
| N | 7665 | hCoV-19/South Africa/KRISP-K003381/2020\|EPI_ISL_60 | ···················· | ·A·················· | ···················· | ···················· | ···················· | ···················· |
| N | 7666 | hCoV-19/South Africa/KRISP-K003382/2020\|EPI_ISL_60 | ···················· | ···················· | ···················· | ···················· | ···················· | ···················· |
| N | 7667 | hCoV-19/South Africa/KRISP-K003383/2020\|EPI_ISL_60 | ···················· | ···················· | ···················· | ···················· | ···················· | ···················· |
| N | 7668 | hCoV-19/South Africa/KRISP-K003384/2020\|EPI_ISL_60 | ···················· | ·A·················· | ···················· | ···················· | ···················· | ···················· |
| N | 7669 | hCoV-19/South Africa/KRISP-K003387/2020\|EPI_ISL_60 | ···················· | ···················· | ···················· | ···················· | ···················· | ···················· |
| N | 7670 | hCoV-19/South Africa/KRISP-K003388/2020\|EPI_ISL_60 | ···················· | ···················· | ···················· | ···················· | ···················· | ···················· |
| N | 7672 | hCoV-19/South Africa/KRISP-K003390/2020\|EPI_ISL_60 | ···················· | ···················· | ···················· | ···················· | ···················· | ···················· |
| N | 7673 | hCoV-19/South Africa/KRISP-K003391/2020\|EPI_ISL_60 | ···················· | ···················· | ···················· | ···················· | ···················· | ···················· |
| N | 7674 | hCoV-19/USA/MN-QDX-1632/2020\|EPI_ISL_603993\|2020-0 | ···················· | ···················· | ···················· | ···················· | ···················· | ···················· |
| N | 7675 | hCoV-19/USA/MN-QDX-1643/2020\|EPI_ISL_603994\|2020-0 | ···················· | ···················· | ···················· | ···················· | ···················· | ···················· |
| N | 7676 | hCoV-19/USA/MN-QDX-1646/2020\|EPI_ISL_603995\|2020-0 | ···················· | ···················· | ···················· | ···················· | ···················· | ···················· |
| N | 7677 | hCoV-19/USA/MN-QDX-1647/2020\|EPI_ISL_603996\|2020-0 | ···················· | ···················· | ···················· | ···················· | ···················· | ···················· |
| N | 7678 | hCoV-19/USA/MN-QDX-1650/2020\|EPI_ISL_603997\|2020-0 | ···················· | ···················· | ···················· | ···················· | ···················· | ···················· |
| N | 7679 | hCoV-19/USA/MN-QDX-1652/2020\|EPI_ISL_603998\|2020-0 | ···················· | ···················· | ···················· | ···················· | ···················· | ···················· |
| N | 7680 | hCoV-19/USA/MN-QDX-1653/2020\|EPI_ISL_603999\|2020-0 | ···················· | ···················· | ···················· | ···················· | ···················· | ···················· |
| N | 7681 | hCoV-19/USA/WA-S2857/2020\|EPI_ISL_596388\|2020-09-1 | ···················· | ···················· | ···················· | ···················· | ···················· | ···················· |
| N | 7682 | hCoV-19/USA/WA-S2858/2020\|EPI_ISL_596389\|2020-09-1 | ···················· | ·············A······ | ···················· | ···················· | ···················· | ···················· |
| N | 7683 | hCoV-19/USA/WA-S2860/2020\|EPI_ISL_596391\|2020-09-1 | ···················· | ·A·················· | ···················· | ···················· | ···················· | ···················· |
| N | 7684 | hCoV-19/USA/WA-S2863/2020\|EPI_ISL_596394\|2020-09-1 | ···················· | ···················· | ···················· | ···················· | ···················· | ···················· |
| N | 7685 | hCoV-19/USA/WA-S2864/2020\|EPI_ISL_596395\|2020-09-1 | ···················· | ···················· | ···················· | ···················· | ···················· | ···················· |
| N | 7686 | hCoV-19/USA/WA-S2865/2020\|EPI_ISL_596396\|2020-09-1 | ···················· | ···················· | ···················· | ···················· | ···················· | ···················· |
| N | 7687 | hCoV-19/USA/WA-S2866/2020\|EPI_ISL_596397\|2020-09-2 | ···················· | ···················· | ···················· | ···················· | ···················· | ···················· |
| N | 7688 | hCoV-19/USA/WA-S2867/2020\|EPI_ISL_596398\|2020-09-2 | ···················· | ···················· | ···················· | ···················· | ···················· | ···················· |
| N | 7689 | hCoV-19/England/ALDP-A50C4F/2020\|EPI_ISL_597612\|20 | ···················· | ···················· | ···················· | ···················· | ···················· | ···················· |
| N | 7690 | hCoV-19/England/ALDP-A50289/2020\|EPI_ISL_597613\|20 | ···················· | ···················· | ···················· | ···················· | ···················· | ···················· |
| N | 7691 | hCoV-19/England/ALDP-A506E7/2020\|EPI_ISL_597614\|20 | ···················· | ···················· | ···················· | ···················· | ···················· | ···················· |
| N | 7692 | hCoV-19/England/ALDP-A50F55/2020\|EPI_ISL_597615\|20 | ···················· | ···················· | ···················· | ···················· | ···················· | ···················· |
| N | 7693 | hCoV-19/England/ALDP-A50465/2020\|EPI_ISL_597616\|20 | ···················· | ···················· | ···················· | ···················· | ···················· | ···················· |
| N | 7694 | hCoV-19/England/ALDP-A50CD6/2020\|EPI_ISL_597617\|20 | ···················· | ···················· | ···················· | ···················· | ···················· | ···················· |
| N | 7695 | hCoV-19/England/ALDP-A50BAC/2020\|EPI_ISL_597618\|20 | ···················· | ·········A·········· | ···················· | ···················· | ···················· | ···················· |
| N | 7696 | hCoV-19/England/ALDP-A508C3/2020\|EPI_ISL_597619\|20 | ···················· | ···················· | ···················· | ···················· | ···················· | ···················· |
| N | 7697 | hCoV-19/England/ALDP-A500BC/2020\|EPI_ISL_597620\|20 | ···················· | ···················· | ···················· | ···················· | ···················· | ···················· |
| N | 7698 | hCoV-19/England/ALDP-A4FE7A/2020\|EPI_ISL_597622\|20 | ···················· | ···················· | ···················· | ···················· | ···················· | ···················· |
| N | 7699 | hCoV-19/England/ALDP-A50DC4/2020\|EPI_ISL_597623\|20 | ···················· | ···················· | ···················· | ···················· | ···················· | ···················· |
| N | 7700 | hCoV-19/England/ALDP-A50711/2020\|EPI_ISL_597624\|20 | ···················· | ···················· | ···················· | ···················· | ···················· | ···················· |
| N | 7701 | hCoV-19/England/ALDP-A4FF95/2020\|EPI_ISL_597625\|20 | ···················· | ···················· | ···················· | ···················· | ···················· | ···················· |
| N | 7702 | hCoV-19/England/ALDP-A50052/2020\|EPI_ISL_597626\|20 | ···················· | ···················· | ···················· | ···················· | ···················· | ···················· |
| N | 7703 | hCoV-19/England/ALDP-A4FEF2/2020\|EPI_ISL_597627\|20 | ···················· | ···················· | ···················· | ···················· | ···················· | ···················· |
| N | 7704 | hCoV-19/England/ALDP-A50CE5/2020\|EPI_ISL_597628\|20 | ···················· | ·A·················· | ···················· | ···················· | ···················· | ···A················ |
| N | 7705 | hCoV-19/England/ALDP-A504FC/2020\|EPI_ISL_597629\|20 | ···················· | ···················· | ···················· | ···················· | ···················· | ···················· |
| N | 7706 | hCoV-19/England/ALDP-A4FBDD/2020\|EPI_ISL_597630\|20 | ···················· | ···················· | ···················· | ···················· | ···················· | ···················· |
| N | 7707 | hCoV-19/England/ALDP-A50A45/2020\|EPI_ISL_597676\|20 | ···················· | ···················· | ···················· | ···················· | ···················· | ···················· |
| N | 7708 | hCoV-19/England/ALDP-A50887/2020\|EPI_ISL_597677\|20 | ···················· | ···················· | ···················· | ···················· | ···················· | ···················· |
| N | 7709 | hCoV-19/England/ALDP-A4FE6B/2020\|EPI_ISL_597678\|20 | ···················· | ···················· | ···················· | ···················· | ···················· | ···················· |
| N | 7710 | hCoV-19/England/ALDP-A4FD31/2020\|EPI_ISL_597679\|20 | ···················· | ···················· | ···················· | ···················· | ···················· | ···················· |
| N | 7711 | hCoV-19/England/ALDP-A50ACD/2020\|EPI_ISL_597680\|20 | ···················· | ···················· | ···················· | ···················· | ···················· | ···················· |
| N | 7712 | hCoV-19/England/ALDP-A4FEA7/2020\|EPI_ISL_597681\|20 | ···················· | ···················· | ···················· | ···················· | ···················· | ···················· |
| N | 7713 | hCoV-19/England/ALDP-A508A5/2020\|EPI_ISL_597682\|20 | ···················· | ···················· | ···················· | ···················· | ···················· | ···················· |
| N | 7714 | hCoV-19/England/ALDP-A4FCE9/2020\|EPI_ISL_597683\|20 | ···················· | ·A·················· | ···················· | ···················· | ···················· | ···A················ |
| N | 7715 | hCoV-19/England/ALDP-A4FDC8/2020\|EPI_ISL_597684\|20 | ···················· | ···················· | ···················· | ···················· | ···················· | ···················· |
| N | 7716 | hCoV-19/England/ALDP-A4FAC1/2020\|EPI_ISL_597685\|20 | ···················· | ···················· | ···················· | ···················· | ···················· | ···················· |
| N | 7717 | hCoV-19/England/ALDP-A5084B/2020\|EPI_ISL_597686\|20 | ···················· | ···················· | ···················· | ···················· | ···················· | ···················· |
| N | 7718 | hCoV-19/England/ALDP-A50702/2020\|EPI_ISL_597687\|20 | ···················· | ···················· | ···················· | ···················· | ···················· | ···················· |
| N | 7719 | hCoV-19/England/ALDP-A5022F/2020\|EPI_ISL_597688\|20 | ···················· | ···················· | ···················· | ···················· | ···················· | ···················· |
| N | 7720 | hCoV-19/England/ALDP-A50632/2020\|EPI_ISL_597689\|20 | ···················· | ·A·················· | ···················· | ···················· | ···················· | ···A················ |
| N | 7721 | hCoV-19/England/ALDP-A5081E/2020\|EPI_ISL_597690\|20 | ···················· | ···················· | ···················· | ···················· | ···················· | ···················· |
| N | 7722 | hCoV-19/England/ALDP-A4FB82/2020\|EPI_ISL_597691\|20 | ···················· | ···················· | ···················· | ···················· | ···················· | ···················· |
| N | 7723 | hCoV-19/England/ALDP-A503FF/2020\|EPI_ISL_597692\|20 | ···················· | ···················· | ···················· | ···················· | ···················· | ···················· |
| N | 7724 | hCoV-19/England/ALDP-A4FE98/2020\|EPI_ISL_597693\|20 | ···················· | ···················· | ···················· | ···················· | ···················· | ···················· |
| N | 7725 | hCoV-19/England/ALDP-A50377/2020\|EPI_ISL_597694\|20 | ···················· | ···················· | ···················· | ···················· | ···················· | ···················· |
| N | 7726 | hCoV-19/England/ALDP-A5027A/2020\|EPI_ISL_597696\|20 | ···················· | ···················· | ···················· | ···················· | ···················· | ···················· |
| N | 7727 | hCoV-19/England/ALDP-A50E94/2020\|EPI_ISL_597697\|20 | ···················· | ···················· | ···················· | ···················· | ···················· | ···················· |
| N | 7728 | hCoV-19/England/ALDP-A506D8/2020\|EPI_ISL_597698\|20 | ···················· | ···················· | ···················· | ···················· | ···················· | ···················· |
| N | 7729 | hCoV-19/England/ALDP-A50E49/2020\|EPI_ISL_597699\|20 | ···················· | ···················· | ···················· | ···················· | ···················· | ···················· |
| N | 7730 | hCoV-19/Scotland/QEUH-A1EE38/2020\|EPI_ISL_598900\|2 | ···················· | ···················· | ···················· | ···················· | ···················· | ···················· |

N 7731 hCoV-19/Scotland/QEUH-A1E09C/2020|EPI_ISL_598901|2 ···················· ···················· ···················· ···················· ···················· ····················
N 7732 hCoV-19/Scotland/QEUH-A217EC/2020|EPI_ISL_598902|2 ···················· ···················· ···················· ···················· ······T············· ····················
N 7733 hCoV-19/Scotland/QEUH-A1E551/2020|EPI_ISL_598903|2 ···················· ···················· ···················· ···················· ···················· ····················
N 7734 hCoV-19/Scotland/QEUH-A1E041/2020|EPI_ISL_598904|2 ···················· ···················· ···················· ···················· ···················· ····················
N 7735 hCoV-19/Scotland/QEUH-A1E31B/2020|EPI_ISL_598910|2 ···················· ···················· ···················· ···················· ···················· ····················
N 7736 hCoV-19/Scotland/QEUH-A1DD78/2020|EPI_ISL_598911|2 ···················· ···················· ···················· ···················· ···················· ····················
N 7737 hCoV-19/Scotland/QEUH-A1DA26/2020|EPI_ISL_598912|2 ···················· ···················· ···················· ···················· ···················· ····················
N 7738 hCoV-19/Scotland/QEUH-A1EA43/2020|EPI_ISL_598913|2 ···················· ···················· ···················· ···················· ···················· ····················
N 7739 hCoV-19/Scotland/QEUH-A20316/2020|EPI_ISL_598914|2 ···················· ···················· ···················· ···················· ···················· ····················
N 7740 hCoV-19/Scotland/QEUH-A1FE55/2020|EPI_ISL_598915|2 ···················· ···················· ···················· ···················· ···················· ····················
N 7741 hCoV-19/Scotland/QEUH-A1F72D/2020|EPI_ISL_598916|2 ···················· ···················· ···················· ···················· ···················· ····················
N 7742 hCoV-19/Scotland/QEUH-A1F59C/2020|EPI_ISL_598917|2 ···················· ···················· ···················· ···················· ···················· ····················
N 7743 hCoV-19/Scotland/QEUH-A1FD67/2020|EPI_ISL_598918|2 ···················· ···················· ···················· ···················· ···················· ····················
N 7744 hCoV-19/Scotland/QEUH-A1FF61/2020|EPI_ISL_598927|2 ···················· ···················· ···················· ···················· ···················· ····················
N 7745 hCoV-19/Scotland/QEUH-A1FE28/2020|EPI_ISL_598928|2 ···················· ···················· ···················· ···················· ···················· ····················
N 7746 hCoV-19/Scotland/QEUH-A1FD1C/2020|EPI_ISL_598941|2 ···················· ···················· ···················· ···················· ···················· ····················
N 7747 hCoV-19/Scotland/QEUH-A20422/2020|EPI_ISL_598942|2 ···················· ···················· ···················· ···················· ···················· ····················
N 7748 hCoV-19/Scotland/QEUH-A1F67B/2020|EPI_ISL_598943|2 ···················· ···················· ···················· ···················· ···················· ····················
N 7749 hCoV-19/Scotland/QEUH-A22217/2020|EPI_ISL_598944|2 ···················· ···················· ···················· ···················· ···················· ····················
N 7750 hCoV-19/Scotland/QEUH-A1F857/2020|EPI_ISL_598946|2 ···················· ···················· ···················· ···················· ···················· ····················
N 7751 hCoV-19/Scotland/QEUH-A1F866/2020|EPI_ISL_598947|2 ···················· ···················· ···················· ···················· ···················· ····················
N 7752 hCoV-19/Scotland/QEUH-A224F3/2020|EPI_ISL_598948|2 ···················· ···················· ···················· ···················· ···················· ····················
N 7753 hCoV-19/Scotland/QEUH-A1FD76/2020|EPI_ISL_598949|2 ···················· ···················· ···················· ···················· ···················· ····················
N 7754 hCoV-19/Scotland/QEUH-A20D27/2020|EPI_ISL_598950|2 ···················· ···················· ···················· ···················· ···················· ····················
N 7755 hCoV-19/Scotland/QEUH-A1FD85/2020|EPI_ISL_598951|2 ···················· ···················· ···················· ···················· ···················· ····················
N 7756 hCoV-19/Scotland/QEUH-A1FEDD/2020|EPI_ISL_598952|2 ···················· ···················· ···················· ···················· ···················· ····················
N 7757 hCoV-19/Scotland/QEUH-A1F8FD/2020|EPI_ISL_598953|2 ···················· ···················· ···················· ···················· ···················· ····················
N 7758 hCoV-19/Scotland/QEUH-A20E24/2020|EPI_ISL_598954|2 ···················· ···················· ···················· ···················· ···················· ····················
N 7759 hCoV-19/Scotland/QEUH-A1FAAC/2020|EPI_ISL_598955|2 ···················· ···················· ···················· ···················· ···················· ····················
N 7760 hCoV-19/Scotland/QEUH-A2070B/2020|EPI_ISL_598956|2 ···················· ···················· ···················· ···················· ···················· ····················
N 7761 hCoV-19/Scotland/QEUH-A1FF43/2020|EPI_ISL_598957|2 ···················· ···················· ···················· ···················· ···················· ····················
N 7762 hCoV-19/Scotland/QEUH-A203DA/2020|EPI_ISL_598958|2 ···················· ···················· ···················· ···················· ···················· ····················
N 7763 hCoV-19/Scotland/QEUH-A1F71E/2020|EPI_ISL_598960|2 ···················· ···················· ···················· ···················· ···················· ····················
N 7764 hCoV-19/Scotland/QEUH-A22727/2020|EPI_ISL_598961|2 ···················· ···················· ···················· ···················· ···················· ····················
N 7765 hCoV-19/Scotland/QEUH-A1FF52/2020|EPI_ISL_598962|2 ···················· ···················· ···················· ···················· ···················· ····················
N 7766 hCoV-19/England/MILK-A056C3/2020|EPI_ISL_600004|20 ···················· ···················· ···················· ···················· ···················· ····················
N 7767 hCoV-19/England/MILK-A054C9/2020|EPI_ISL_600008|20 ···················· ···················· ···················· ···················· ···················· ····················
N 7768 hCoV-19/England/MILK-A06880/2020|EPI_ISL_600009|20 ···················· ···················· ···················· ···················· ···················· ····················
N 7769 hCoV-19/England/MILK-A057C0/2020|EPI_ISL_600010|20 ···················· ·A·················· ···················· ···················· ···················· ··A·················
N 7770 hCoV-19/England/MILK-A053EA/2020|EPI_ISL_600011|20 ···················· ···················· ···················· ···················· ···················· ····················
N 7771 hCoV-19/England/MILK-A05380/2020|EPI_ISL_600012|20 ···················· ···················· ···················· ···················· ···················· ····················
N 7772 hCoV-19/England/MILK-A066E0/2020|EPI_ISL_600013|20 ···················· ···················· ···················· ···················· ······T············· ····················
N 7773 hCoV-19/England/MILK-A05E43/2020|EPI_ISL_600014|20 ···················· ·A·················· ···················· ···················· ···················· ··A·················
N 7774 hCoV-19/England/MILK-A05A03/2020|EPI_ISL_600015|20 ···················· ···················· ···················· ···················· ···················· ····················
N 7775 hCoV-19/England/MILK-A05C1C/2020|EPI_ISL_600016|20 ···················· ···················· ···················· ···················· ···················· ····················
N 7776 hCoV-19/England/MILK-A05D0A/2020|EPI_ISL_600017|20 ···················· ···················· ···················· ···················· ···················· ····················
N 7777 hCoV-19/England/MILK-A060D3/2020|EPI_ISL_600018|20 ···················· ···················· ···················· ···················· ···················· ····················
N 7778 hCoV-19/Wales/MILK-A063CB/2020|EPI_ISL_600019|2020 ···················· ···················· ···················· ···················· ···················· ····················
N 7779 hCoV-19/England/MILK-A05915/2020|EPI_ISL_600020|20 ···················· ···················· ···················· ···················· ···················· ····················
N 7780 hCoV-19/England/MILK-A0653E/2020|EPI_ISL_600021|20 ···················· ···················· ···················· ···················· ···················· ····················
N 7781 hCoV-19/England/MILK-A05AE5/2020|EPI_ISL_600022|20 ···················· ···················· ···················· ···················· ···················· ····················
N 7782 hCoV-19/England/MILK-A0604C/2020|EPI_ISL_600023|20 ···················· ···················· ···················· ···················· ···················· ····················
N 7783 hCoV-19/England/MILK-A0561E/2020|EPI_ISL_600024|20 ···················· ···················· ···················· ···················· ···················· ····················
N 7784 hCoV-19/England/MILK-A053F9/2020|EPI_ISL_600025|20 ···················· ···················· ···················· ···················· ···················· ····················
N 7785 hCoV-19/England/MILK-A06747/2020|EPI_ISL_600026|20 ···················· ···················· ···················· ···················· ···················· ····················
N 7786 hCoV-19/England/MILK-A0639E/2020|EPI_ISL_600027|20 ···················· ···················· ···················· ···················· ···················· ····················
N 7787 hCoV-19/England/MILK-A061A3/2020|EPI_ISL_600028|20 ···················· ···················· ···················· ···················· ···················· ····················
N 7788 hCoV-19/England/MILK-A06862/2020|EPI_ISL_600029|20 ···················· ···················· ···················· ···················· ···················· ····················
N 7789 hCoV-19/England/MILK-A05D28/2020|EPI_ISL_600030|20 ···················· ···················· ···················· ···················· ···················· ····················
N 7790 hCoV-19/England/MILK-A06282/2020|EPI_ISL_600031|20 ···················· ···················· ···················· ···················· ···················· ····················
N 7791 hCoV-19/England/MILK-A05784/2020|EPI_ISL_600032|20 ···················· ·A·················· ···················· ···················· ···················· ··A·················
N 7792 hCoV-19/England/MILK-A05B79/2020|EPI_ISL_600034|20 ···················· ·A·················· ···················· ···················· ···················· ····················
N 7793 hCoV-19/England/MILK-A06808/2020|EPI_ISL_600035|20 ···················· ···················· ···················· ···················· ···················· ····················
N 7794 hCoV-19/England/MILK-A0565A/2020|EPI_ISL_600036|20 ···················· ···················· ···················· ···················· ···················· ····················
N 7795 hCoV-19/England/MILK-A05432/2020|EPI_ISL_600037|20 ···················· ···················· ···················· ···················· ···················· ····················
N 7796 hCoV-19/England/MILK-A05B88/2020|EPI_ISL_600038|20 ···················· ···················· ···················· ···················· ···················· ····················
N 7797 hCoV-19/England/MILK-A05BD3/2020|EPI_ISL_600039|20 ···················· ···················· ···················· ···················· ···················· ····················
N 7798 hCoV-19/England/MILK-A0660E/2020|EPI_ISL_600080|20 ···················· ···················· ···················· ···················· ······T············· ····················
N 7799 hCoV-19/England/MILK-A06316/2020|EPI_ISL_600081|20 ···················· ···················· ···················· ···················· ···················· ····················
N 7800 hCoV-19/England/MILK-A0647D/2020|EPI_ISL_600082|20 ···················· ·A·················· ···················· ···················· ···················· ····················
N 7801 hCoV-19/England/MILK-A05942/2020|EPI_ISL_600084|20 ···················· ···················· ···················· ···················· ···················· ····················
N 7802 hCoV-19/England/MILK-A06097/2020|EPI_ISL_600085|20 ···················· ···················· ···················· ···················· ···················· ····················
N 7803 hCoV-19/England/MILK-A067DE/2020|EPI_ISL_600086|20 ···················· ···················· ···················· ···················· ···················· ····················
N 7804 hCoV-19/England/MILK-A054F6/2020|EPI_ISL_600087|20 ···················· ···················· ···················· ···················· ···················· ····················
N 7805 hCoV-19/Wales/MILK-A069E7/2020|EPI_ISL_600088|2020 ···················· ···················· ···················· ···················· ···················· ····················
N 7806 hCoV-19/England/MILK-A05F22/2020|EPI_ISL_600089|20 ···················· ···················· ···················· ···················· ···················· ····················
N 7807 hCoV-19/England/MILK-A055D5/2020|EPI_ISL_600090|20 ···················· ···················· ···················· ···················· ···················· ····················
N 7808 hCoV-19/England/MILK-A05511/2020|EPI_ISL_600094|20 ···················· ···················· ···················· ···················· ···················· ····················
N 7809 hCoV-19/England/MILK-A0606A/2020|EPI_ISL_600095|20 ···················· ···················· ···················· ···················· ···················· ····················
N 7810 hCoV-19/England/MILK-A0546F/2020|EPI_ISL_600096|20 ···················· ···················· ···················· ···················· ···················· ····················

| | | | | | | | | |
|---|---|---|---|---|---|---|---|---|
| N | 7811 hCoV-19/England/MILK-A06325/2020\|EPI_ISL_600097\|20 | ···················· | ···················· | ···················· | ···················· | ···················· | ···················· | |
| N | 7812 hCoV-19/England/MILK-A0663B/2020\|EPI_ISL_600098\|20 | ···················· | ···················· | ···················· | ···················· | ···················· | ···················· | |
| N | 7813 hCoV-19/England/MILK-A066A4/2020\|EPI_ISL_600099\|20 | ···················· | ···················· | ···················· | ···················· | ···················· | ···················· | |
| N | 7814 hCoV-19/England/QEUH-9DBD86/2020\|EPI_ISL_601300\|20 | ···················· | ·A·················· | ···················· | ···················· | ···················· | ··A················· | |
| N | 7815 hCoV-19/England/MILK-9E046B/2020\|EPI_ISL_601302\|20 | ···················· | ···················· | ···················· | ···················· | ···················· | ···················· | |
| N | 7816 hCoV-19/England/MILK-9E08D8/2020\|EPI_ISL_601303\|20 | ···················· | ···················· | ···················· | ···················· | ···················· | ···················· | |
| N | 7817 hCoV-19/England/MILK-9E0B39/2020\|EPI_ISL_601304\|20 | ···················· | ···················· | ···················· | ···················· | ···················· | ···················· | |
| N | 7818 hCoV-19/England/MILK-9E06CF/2020\|EPI_ISL_601305\|20 | ···················· | ···················· | ···················· | ···················· | ···················· | ···················· | |
| N | 7819 hCoV-19/Wales/MILK-9E0A69/2020\|EPI_ISL_601306\|2020 | ···················· | ···················· | ···················· | ···················· | ···················· | ···················· | |
| N | 7820 hCoV-19/England/MILK-9E0B93/2020\|EPI_ISL_601307\|20 | ···················· | ···················· | ···················· | ···················· | ···················· | ···················· | |
| N | 7821 hCoV-19/England/QEUH-9DA8D1/2020\|EPI_ISL_601308\|20 | ···················· | ···················· | ···················· | ···················· | ···················· | ···················· | |
| N | 7822 hCoV-19/England/QEUH-9DC2B3/2020\|EPI_ISL_601309\|20 | ···················· | ·A·················· | ···················· | ···················· | ···················· | ··A················· | |
| N | 7823 hCoV-19/Wales/MILK-9E0823/2020\|EPI_ISL_601311\|2020 | ···················· | ···················· | ···················· | ···················· | ···················· | ···················· | |
| N | 7824 hCoV-19/Wales/MILK-9E050E/2020\|EPI_ISL_601312\|2020 | ···················· | ···················· | ···················· | ···················· | ···················· | ···················· | |
| N | 7825 hCoV-19/England/MILK-9E02CB/2020\|EPI_ISL_601330\|20 | ···················· | ···················· | ···················· | ···················· | ···················· | ···················· | |
| N | 7826 hCoV-19/England/QEUH-9DC0B9/2020\|EPI_ISL_601335\|20 | ···················· | ···················· | ···················· | ···················· | ···················· | ···················· | |
| N | 7827 hCoV-19/England/QEUH-9DB023/2020\|EPI_ISL_601339\|20 | ···················· | ·A·················· | ···················· | ···················· | ···················· | ··A················· | |
| N | 7828 hCoV-19/England/MILK-9E0340/2020\|EPI_ISL_601340\|20 | ···················· | ···················· | ···················· | ···················· | ···················· | ···················· | |
| N | 7829 hCoV-19/England/QEUH-9DA5AD/2020\|EPI_ISL_601341\|20 | ···················· | ·A·················· | ···················· | ···················· | ···················· | ···················· | |
| N | 7830 hCoV-19/England/QEUH-9DC2A4/2020\|EPI_ISL_601342\|20 | ···················· | ···················· | ···················· | ···················· | ···················· | ···················· | |
| N | 7831 hCoV-19/England/MILK-9E03D7/2020\|EPI_ISL_601343\|20 | ···················· | ···················· | ···················· | ···················· | ···················· | ···················· | |
| N | 7832 hCoV-19/England/QEUH-9DA5F8/2020\|EPI_ISL_601344\|20 | ···················· | ·A·················· | ···················· | ···················· | ···················· | ···················· | |
| N | 7833 hCoV-19/England/MILK-9E054A/2020\|EPI_ISL_601345\|20 | ···················· | ···················· | ···················· | ···················· | ···················· | ···················· | |
| N | 7834 hCoV-19/Wales/MILK-9E060B/2020\|EPI_ISL_601346\|2020 | ···················· | ···················· | ···················· | ···················· | ···················· | ···················· | |
| N | 7835 hCoV-19/England/QEUH-9DA6AA/2020\|EPI_ISL_601347\|20 | ···················· | ···················· | ···················· | ···················· | ······T············· | ···················· | |
| N | 7836 hCoV-19/England/QEUH-9DA8B3/2020\|EPI_ISL_601348\|20 | ···················· | ···················· | ···················· | ···················· | ···················· | ···················· | |
| N | 7837 hCoV-19/England/MILK-9E0BC0/2020\|EPI_ISL_601349\|20 | ···················· | ···················· | ···················· | ···················· | ······T············· | ···················· | |
| N | 7838 hCoV-19/Wales/MILK-9E0735/2020\|EPI_ISL_601350\|2020 | ···················· | ···················· | ···················· | ···················· | ···················· | ···················· | |
| N | 7839 hCoV-19/England/QEUH-9DA525/2020\|EPI_ISL_601351\|20 | ···················· | ·A·················· | ···················· | ···················· | ···················· | ···················· | |
| N | 7840 hCoV-19/England/QEUH-9DBE83/2020\|EPI_ISL_601352\|20 | ···················· | ···················· | ···················· | ···················· | ···················· | ···················· | |
| N | 7841 hCoV-19/England/QEUH-9DBECF/2020\|EPI_ISL_601353\|20 | ···················· | ···················· | ···················· | ···················· | ···················· | ···················· | |
| N | 7842 hCoV-19/England/MILK-9E0A96/2020\|EPI_ISL_601354\|20 | ···················· | ···················· | ···················· | ···················· | ···················· | ···················· | |
| N | 7843 hCoV-19/England/QEUH-9DC004/2020\|EPI_ISL_601355\|20 | ···················· | ···················· | ···················· | ···················· | ···················· | ···················· | |
| N | 7844 hCoV-19/England/QEUH-9DA570/2020\|EPI_ISL_601356\|20 | ···················· | ···················· | ···················· | ···················· | ···················· | ···················· | |
| N | 7845 hCoV-19/England/QEUH-9DBEFC/2020\|EPI_ISL_601357\|20 | ···················· | ···················· | ···················· | ···················· | ···················· | ···················· | |
| N | 7846 hCoV-19/England/QEUH-9DA8E0/2020\|EPI_ISL_601358\|20 | ···················· | ···················· | ···················· | ···················· | ···················· | ···················· | |
| N | 7847 hCoV-19/England/MILK-9E07AE/2020\|EPI_ISL_601359\|20 | ···················· | ···················· | ···················· | ···················· | ···················· | ···················· | |
| N | 7848 hCoV-19/England/MILK-9E0647/2020\|EPI_ISL_601360\|20 | ···················· | ···················· | ···················· | ···················· | ···················· | ···················· | |
| N | 7849 hCoV-19/England/QEUH-9DA76B/2020\|EPI_ISL_601362\|20 | ···················· | ···················· | ···················· | ···················· | ···················· | ···················· | |
| N | 7850 hCoV-19/England/MILK-9E0322/2020\|EPI_ISL_601363\|20 | ···················· | ···················· | ···················· | ···················· | ···················· | ···················· | |
| N | 7851 hCoV-19/England/QEUH-9DA65F/2020\|EPI_ISL_601364\|20 | ···················· | ···················· | ···················· | ···················· | ···················· | ···················· | |
| N | 7852 hCoV-19/England/MILK-9E02E9/2020\|EPI_ISL_601365\|20 | ···················· | ···················· | ···················· | ···················· | ···················· | ···················· | |
| N | 7853 hCoV-19/Wales/MILK-9E0C72/2020\|EPI_ISL_601367\|2020 | ···················· | ···················· | ···················· | ···················· | ···················· | ···················· | |
| N | 7854 hCoV-19/England/MILK-9E028F/2020\|EPI_ISL_601368\|20 | ···················· | ···················· | ···················· | ···················· | ···················· | ···················· | |
| N | 7855 hCoV-19/England/MILK-9E0638/2020\|EPI_ISL_601369\|20 | ···················· | ···················· | ···················· | ···················· | ···················· | ···················· | |
| N | 7856 hCoV-19/South Africa/KRISP-K002868/2020\|EPI_ISL_60 | ···················· | ···················· | ···················· | ···················· | ···················· | ···················· | |
| N | 7857 hCoV-19/South Africa/KRISP-K003668/2020\|EPI_ISL_60 | ···················· | ···················· | ···················· | ···················· | ···················· | ···················· | |
| N | 7859 hCoV-19/South Africa/KRISP-K003670/2020\|EPI_ISL_60 | ···················· | ···················· | ···················· | ···················· | ···················· | ···················· | |
| N | 7860 hCoV-19/South Africa/KRISP-K003671/2020\|EPI_ISL_60 | ···················· | ···················· | ···················· | ···················· | ···················· | ···················· | |
| N | 7861 hCoV-19/South Africa/KRISP-K003672/2020\|EPI_ISL_60 | ···················· | ···················· | ···················· | ···················· | ···················· | ···················· | |
| N | 7862 hCoV-19/South Africa/KRISP-K003673/2020\|EPI_ISL_60 | ···················· | ···················· | ···················· | ···················· | ···················· | ···················· | |
| N | 7863 hCoV-19/South Africa/KRISP-K003674/2020\|EPI_ISL_60 | ···················· | ···················· | ···················· | ···················· | ···················· | ···················· | |
| N | 7864 hCoV-19/South Africa/KRISP-K002810/2020\|EPI_ISL_60 | ···················· | ···················· | ···················· | ···················· | ···················· | ···················· | |
| N | 7865 hCoV-19/South Africa/KRISP-K002812/2020\|EPI_ISL_60 | ···················· | ···················· | ···················· | ···················· | ···················· | ···················· | |
| N | 7866 hCoV-19/South Africa/KRISP-K002813/2020\|EPI_ISL_60 | ···················· | ···················· | ···················· | ···················· | ···················· | ···················· | |
| N | 7867 hCoV-19/South Africa/KRISP-K002814/2020\|EPI_ISL_60 | ···················· | ···················· | ···················· | ···················· | ···················· | ···················· | |
| N | 7868 hCoV-19/South Africa/KRISP-K002815/2020\|EPI_ISL_60 | ···················· | ···················· | ···················· | ···················· | ···················· | ···················· | |
| N | 7869 hCoV-19/South Africa/KRISP-K002816/2020\|EPI_ISL_60 | ···················· | ···················· | ···················· | ···················· | ···················· | ···················· | |
| N | 7870 hCoV-19/South Africa/KRISP-K002817/2020\|EPI_ISL_60 | ···················· | ···················· | ···················· | ···················· | ···················· | ···················· | |
| N | 7871 hCoV-19/South Africa/KRISP-K002818/2020\|EPI_ISL_60 | ···················· | ·A·················· | ···················· | ···················· | ···················· | ···················· | |
| N | 7872 hCoV-19/South Africa/KRISP-K002824/2020\|EPI_ISL_60 | ···················· | ···················· | ···················· | ···················· | ···················· | ···················· | |
| N | 7873 hCoV-19/South Africa/KRISP-K002825/2020\|EPI_ISL_60 | ···················· | ···················· | ···················· | ···················· | NNN················· | ···················· | 1 |
| N | 7874 hCoV-19/South Africa/KRISP-K002844/2020\|EPI_ISL_60 | ···················· | ···················· | ···················· | ···················· | ···················· | ···················· | |
| N | 7875 hCoV-19/South Africa/KRISP-K002845/2020\|EPI_ISL_60 | ···················· | ···················· | ···················· | ···················· | ···················· | ···················· | |
| N | 7876 hCoV-19/South Africa/KRISP-K002854/2020\|EPI_ISL_60 | ···················· | ···················· | ···················· | ···················· | ···················· | ···················· | |
| N | 7877 hCoV-19/South Africa/KRISP-K002856/2020\|EPI_ISL_60 | ···················· | ···················· | ···················· | ···················· | ···················· | ···················· | |
| N | 7878 hCoV-19/South Africa/KRISP-K002857/2020\|EPI_ISL_60 | ···················· | ···················· | ···················· | ···················· | ···················· | ···················· | |
| N | 7879 hCoV-19/South Africa/KRISP-K002858/2020\|EPI_ISL_60 | ···················· | ···················· | ···················· | ···················· | ···················· | ···················· | |
| N | 7880 hCoV-19/South Africa/KRISP-K002859/2020\|EPI_ISL_60 | ···················· | ···················· | ···················· | ···················· | ···················· | ···················· | |
| N | 7881 hCoV-19/South Africa/KRISP-K002860/2020\|EPI_ISL_60 | ···················· | ···················· | ···················· | ···················· | ···················· | ···················· | |
| N | 7882 hCoV-19/South Africa/KRISP-K002862/2020\|EPI_ISL_60 | ···················· | ···················· | ···················· | ···················· | ···················· | ···················· | |
| N | 7883 hCoV-19/South Africa/KRISP-K002863/2020\|EPI_ISL_60 | ···················· | ···················· | ···················· | ···················· | ···················· | ···················· | |
| N | 7884 hCoV-19/South Africa/KRISP-K002864/2020\|EPI_ISL_60 | ···················· | ···················· | ···················· | ···················· | ···················· | ···················· | |
| N | 7885 hCoV-19/South Africa/KRISP-K002865/2020\|EPI_ISL_60 | ···················· | ·A·················· | ···················· | ···················· | ···················· | ···················· | |
| N | 7886 hCoV-19/South Africa/KRISP-K002866/2020\|EPI_ISL_60 | ···················· | ···················· | ···················· | ···················· | ···················· | ···················· | |
| N | 7887 hCoV-19/South Africa/KRISP-K002867/2020\|EPI_ISL_60 | ···················· | ···················· | ···················· | ···················· | ···················· | ···················· | |
| N | 7888 hCoV-19/South Africa/KRISP-K002869/2020\|EPI_ISL_60 | ···················· | ···················· | ···················· | ···················· | ···················· | ···················· | |
| N | 7889 hCoV-19/South Africa/KRISP-K002870/2020\|EPI_ISL_60 | ···················· | ···················· | ···················· | ···················· | ···················· | ···················· | |
| N | 7890 hCoV-19/South Africa/KRISP-K002871/2020\|EPI_ISL_60 | ···················· | ···················· | ···················· | ···················· | ···················· | ···················· | |
| N | 7891 hCoV-19/South Africa/KRISP-K002872/2020\|EPI_ISL_60 | ···················· | ·A·················· | ···················· | ···················· | ···················· | ···················· | |

| | | | | | | | | |
|---|---|---|---|---|---|---|---|---|
| N | 7892 | hCoV-19/South Africa/KRISP-K002873/2020\|EPI_ISL_60 | ···················· | ···················· | ···················· | ···················· | ···················· | ···················· |
| N | 7893 | hCoV-19/South Africa/KRISP-K002874/2020\|EPI_ISL_60 | ···················· | ·A·················· | ···················· | ···················· | ···················· | ···················· |
| N | 7894 | hCoV-19/South Africa/KRISP-K002875/2020\|EPI_ISL_60 | ···················· | ·A·················· | ···················· | ···················· | ···················· | ···················· |
| N | 7895 | hCoV-19/South Africa/KRISP-K002876/2020\|EPI_ISL_60 | ···················· | ·A·················· | ···················· | ···················· | ···················· | ···················· |
| N | 7896 | hCoV-19/South Africa/KRISP-K002877/2020\|EPI_ISL_60 | ···················· | ·A·················· | ···················· | ···················· | ···················· | ···················· |
| N | 7897 | hCoV-19/South Africa/KRISP-K002878/2020\|EPI_ISL_60 | ···················· | ·A·················· | ···················· | ···················· | ···················· | ···················· |
| N | 7898 | hCoV-19/South Africa/KRISP-K002880/2020\|EPI_ISL_60 | ···················· | ···················· | ···················· | ···················· | ···················· | ···················· |
| N | 7899 | hCoV-19/South Africa/KRISP-K002881/2020\|EPI_ISL_60 | ···················· | ···················· | ···················· | ···················· | ···················· | ···················· |
| N | 7900 | hCoV-19/South Africa/KRISP-K002882/2020\|EPI_ISL_60 | ···················· | ·A·················· | ···················· | ···················· | ···················· | ···················· |
| N | 7901 | hCoV-19/South Africa/KRISP-K002883/2020\|EPI_ISL_60 | ···················· | ···················· | ···················· | ···················· | ···················· | ···················· |
| N | 7902 | hCoV-19/South Africa/KRISP-K002884/2020\|EPI_ISL_60 | ···················· | ···················· | ···················· | ···················· | ···················· | ···················· |
| N | 7903 | hCoV-19/South Africa/KRISP-K002885/2020\|EPI_ISL_60 | ···················· | ···················· | ···················· | ···················· | ···················· | ···················· |
| N | 7904 | hCoV-19/South Africa/KRISP-K002886/2020\|EPI_ISL_60 | ···················· | ···················· | ···················· | ···················· | ···················· | ···················· |
| N | 7905 | hCoV-19/USA/CA-QDX-1599/2020\|EPI_ISL_603900\|2020-0 | ···················· | ···················· | ···················· | ···················· | ···················· | ···················· |
| N | 7906 | hCoV-19/USA/CA-QDX-1605/2020\|EPI_ISL_603901\|2020-0 | ···················· | ···················· | ···················· | ···················· | ···················· | ···················· |
| N | 7907 | hCoV-19/USA/MI-QDX-1564/2020\|EPI_ISL_603946\|2020-0 | ···················· | ···················· | ···················· | ···················· | ···················· | ···················· |
| N | 7908 | hCoV-19/USA/MI-QDX-1570/2020\|EPI_ISL_603947\|2020-0 | ···················· | ···················· | ···················· | ···················· | ···················· | ···················· |
| N | 7909 | hCoV-19/USA/MI-QDX-1573/2020\|EPI_ISL_603948\|2020-0 | ···················· | ···················· | ···················· | ···················· | ···················· | ···················· |
| N | 7910 | hCoV-19/USA/MI-QDX-1574/2020\|EPI_ISL_603949\|2020-0 | ···················· | ···················· | ···················· | ···················· | ···················· | ···················· |
| N | 7911 | hCoV-19/USA/MI-QDX-1588/2020\|EPI_ISL_603950\|2020-0 | ···················· | ···················· | ···················· | ···················· | ···················· | ···················· |
| N | 7912 | hCoV-19/USA/MI-QDX-1591/2020\|EPI_ISL_603951\|2020-0 | ···················· | ···················· | ···················· | ···················· | ···················· | ···················· |
| N | 7913 | hCoV-19/USA/MI-QDX-1592/2020\|EPI_ISL_603952\|2020-0 | ···················· | ···················· | ···················· | ···················· | ···················· | ···················· |
| N | 7914 | hCoV-19/USA/MI-QDX-1594/2020\|EPI_ISL_603953\|2020-0 | ···················· | ···················· | ···················· | ···················· | ···················· | ···················· |
| N | 7915 | hCoV-19/USA/MI-QDX-1595/2020\|EPI_ISL_603954\|2020-0 | ···················· | ···················· | ···················· | ···················· | ···················· | ···················· |
| N | 7916 | hCoV-19/USA/MI-QDX-1596/2020\|EPI_ISL_603955\|2020-0 | ···················· | ···················· | ···················· | ···················· | ···················· | ···················· |
| N | 7917 | hCoV-19/USA/MI-QDX-1600/2020\|EPI_ISL_603956\|2020-0 | ···················· | ···················· | ···················· | ···················· | ···················· | ···················· |
| N | 7918 | hCoV-19/USA/MI-QDX-1612/2020\|EPI_ISL_603957\|2020-0 | ···················· | ···················· | ···················· | ···················· | ···················· | ···················· |
| N | 7919 | hCoV-19/USA/MI-QDX-1613/2020\|EPI_ISL_603958\|2020-0 | ···················· | ···················· | ···················· | ···················· | ···················· | ···················· |
| N | 7920 | hCoV-19/USA/MI-QDX-1614/2020\|EPI_ISL_603959\|2020-0 | ···················· | ···················· | ···················· | ···················· | ···················· | ···················· |
| N | 7921 | hCoV-19/USA/MI-QDX-1627/2020\|EPI_ISL_603960\|2020-0 | ···················· | ···················· | ···················· | ···················· | ···················· | ···················· |
| N | 7922 | hCoV-19/USA/MI-QDX-1630/2020\|EPI_ISL_603961\|2020-0 | ···················· | ···················· | ···················· | ···················· | ···················· | ···················· |
| N | 7923 | hCoV-19/USA/MI-QDX-1635/2020\|EPI_ISL_603962\|2020-0 | ···················· | ···················· | ···················· | ···················· | ···················· | ···················· |
| N | 7924 | hCoV-19/USA/MI-QDX-1636/2020\|EPI_ISL_603963\|2020-0 | ···················· | ···················· | ···················· | ···················· | ···················· | ···················· |
| N | 7925 | hCoV-19/USA/MI-QDX-1637/2020\|EPI_ISL_603964\|2020-0 | ···················· | ···················· | ···················· | ···················· | ···················· | ···················· |
| N | 7926 | hCoV-19/USA/MI-QDX-1639/2020\|EPI_ISL_603965\|2020-0 | ···················· | ···················· | ···················· | ···················· | ···················· | ···················· |
| N | 7927 | hCoV-19/USA/MI-QDX-1649/2020\|EPI_ISL_603966\|2020-0 | ···················· | ···················· | ···················· | ···················· | ···················· | ···················· |
| N | 7928 | hCoV-19/USA/MI-QDX-1665/2020\|EPI_ISL_603967\|2020-0 | ···················· | ···················· | ···················· | ···················· | ···················· | ···················· |
| N | 7929 | hCoV-19/USA/MI-QDX-1672/2020\|EPI_ISL_603968\|2020-0 | ···················· | ···················· | ···················· | ···················· | ···················· | ···················· |
| N | 7930 | hCoV-19/USA/MI-QDX-1681/2020\|EPI_ISL_603969\|2020-0 | ···················· | ···················· | ···················· | ···················· | ···················· | ···················· |
| N | 7931 | hCoV-19/USA/MN-QDX-1565/2020\|EPI_ISL_603975\|2020-0 | ···················· | ···················· | ···················· | ···················· | ···················· | ···················· |
| N | 7932 | hCoV-19/USA/MN-QDX-1575/2020\|EPI_ISL_603976\|2020-0 | ···················· | ···················· | ···················· | ···················· | ···················· | ···················· |
| N | 7933 | hCoV-19/USA/MN-QDX-1581/2020\|EPI_ISL_603977\|2020-0 | ···················· | ···················· | ···················· | ···················· | ···················· | ···················· |
| N | 7934 | hCoV-19/USA/MN-QDX-1619/2020\|EPI_ISL_603990\|2020-0 | ···················· | ···················· | ···················· | ···················· | ···················· | ···················· |
| N | 7935 | hCoV-19/USA/MN-QDX-1621/2020\|EPI_ISL_603991\|2020-0 | ···················· | ···················· | ···················· | ···················· | ···················· | ···················· |
| N | 7936 | hCoV-19/USA/MN-QDX-1631/2020\|EPI_ISL_603992\|2020-0 | ···················· | ···················· | ···················· | ···················· | ···················· | ···················· |
| N | 7937 | hCoV-19/England/ALDP-A505AE/2020\|EPI_ISL_597600\|20 | ···················· | ···················· | ···················· | ···················· | ···················· | ···················· |
| N | 7938 | hCoV-19/England/ALDP-A50F91/2020\|EPI_ISL_597601\|20 | ···················· | ···················· | ···················· | ···················· | ···················· | ···················· |
| N | 7939 | hCoV-19/England/ALDP-A4F997/2020\|EPI_ISL_597602\|20 | ···················· | ···················· | ···················· | ···················· | ···················· | ···················· |
| N | 7940 | hCoV-19/England/ALDP-A50E3A/2020\|EPI_ISL_597603\|20 | ···················· | ···················· | ···················· | ···················· | ···················· | ···················· |
| N | 7941 | hCoV-19/England/ALDP-A501B9/2020\|EPI_ISL_597604\|20 | ···················· | ···················· | ···················· | ···················· | ···················· | ···················· |
| N | 7942 | hCoV-19/England/ALDP-A4F9B5/2020\|EPI_ISL_597605\|20 | ···················· | ···················· | ···················· | ···················· | ···················· | ···················· |
| N | 7943 | hCoV-19/England/ALDP-A4F979/2020\|EPI_ISL_597606\|20 | ···················· | ·A·················· | ···················· | ···················· | ···················· | ···················· |
| N | 7944 | hCoV-19/England/ALDP-A502A7/2020\|EPI_ISL_597607\|20 | ···················· | ···················· | ···················· | ···················· | ···················· | ···················· |
| N | 7945 | hCoV-19/England/ALDP-A503E0/2020\|EPI_ISL_597608\|20 | ···················· | ···················· | ···················· | ···················· | ···················· | ···················· |
| N | 7946 | hCoV-19/England/ALDP-A50FDD/2020\|EPI_ISL_597609\|20 | ···················· | ···················· | ···················· | ···················· | ···················· | ···················· |
| N | 7947 | hCoV-19/England/ALDP-A50641/2020\|EPI_ISL_597632\|20 | ···················· | ···················· | ···················· | ···················· | ···················· | ···················· |
| N | 7948 | hCoV-19/England/ALDP-A50F64/2020\|EPI_ISL_597635\|20 | ···················· | ···················· | ···················· | ···················· | ···················· | ···················· |
| N | 7949 | hCoV-19/England/ALDP-A50D4C/2020\|EPI_ISL_597636\|20 | ···················· | ···················· | ···················· | ···················· | ···················· | ···················· |
| N | 7950 | hCoV-19/England/ALDP-A4FB19/2020\|EPI_ISL_597637\|20 | ···················· | ···················· | ···················· | ···················· | ···················· | ···················· |
| N | 7951 | hCoV-19/England/ALDP-A4FD5F/2020\|EPI_ISL_597638\|20 | ···················· | ···················· | ···················· | ···················· | ···················· | ···················· |
| N | 7952 | hCoV-19/England/ALDP-A5034A/2020\|EPI_ISL_597639\|20 | ···················· | ···················· | ···················· | ···················· | ···················· | ···················· |
| N | 7953 | hCoV-19/England/ALDP-A4FBA0/2020\|EPI_ISL_597640\|20 | ···················· | ·A·················· | ···················· | ···················· | ···················· | ···A················ |
| N | 7954 | hCoV-19/England/ALDP-A50201/2020\|EPI_ISL_597641\|20 | ···················· | ···················· | ···················· | ···················· | ···················· | ···················· |
| N | 7955 | hCoV-19/England/ALDP-A50C30/2020\|EPI_ISL_597642\|20 | ···················· | ···················· | ···················· | ···················· | ···················· | ···················· |
| N | 7956 | hCoV-19/England/ALDP-A50E58/2020\|EPI_ISL_597643\|20 | ···················· | ···················· | ···················· | ···················· | ···················· | ···················· |
| N | 7957 | hCoV-19/England/ALDP-A50DD3/2020\|EPI_ISL_597644\|20 | ···················· | ···················· | ···················· | ···················· | ···················· | ···················· |
| N | 7958 | hCoV-19/England/ALDP-A50650/2020\|EPI_ISL_597645\|20 | ···················· | ···················· | ···················· | ···················· | ···················· | ···················· |
| N | 7959 | hCoV-19/England/ALDP-A4FD6E/2020\|EPI_ISL_597646\|20 | ···················· | ·A·······A·········· | ···················· | ···················· | ···················· | ···················· |
| N | 7960 | hCoV-19/England/ALDP-A5016E/2020\|EPI_ISL_597647\|20 | ···················· | ···················· | ···················· | ···················· | ····T··············· | ···················· |
| N | 7961 | hCoV-19/England/ALDP-A50E85/2020\|EPI_ISL_597648\|20 | ···················· | ··············C····· | ···················· | ···················· | ···················· | ···················· |
| N | 7962 | hCoV-19/England/ALDP-A50D3D/2020\|EPI_ISL_597649\|20 | ···················· | ···················· | ···················· | ···················· | ···················· | ···················· |
| N | 7963 | hCoV-19/England/ALDP-A50483/2020\|EPI_ISL_597650\|20 | ···················· | ···················· | ···················· | ···················· | ···················· | ···················· |
| N | 7964 | hCoV-19/England/ALDP-A4FC43/2020\|EPI_ISL_597651\|20 | ···················· | ···················· | ···················· | ···················· | ···················· | ···················· |
| N | 7965 | hCoV-19/England/ALDP-A503D1/2020\|EPI_ISL_597652\|20 | ···················· | ···················· | ···················· | ···················· | ···················· | ···················· |
| N | 7966 | hCoV-19/England/ALDP-A4FBCE/2020\|EPI_ISL_597653\|20 | ···················· | ···················· | ···················· | ···················· | ···················· | ···················· |
| N | 7967 | hCoV-19/England/ALDP-A50614/2020\|EPI_ISL_597654\|20 | ···················· | ···················· | ···················· | ···················· | ···················· | ···················· |
| N | 7968 | hCoV-19/England/ALDP-A503C2/2020\|EPI_ISL_597655\|20 | ···················· | ···················· | ···················· | ···················· | ···················· | ···················· |
| N | 7969 | hCoV-19/England/ALDP-A4FFFF/2020\|EPI_ISL_597656\|20 | ···················· | ·A·················· | ···················· | ···················· | ···················· | ···················· |
| N | 7970 | hCoV-19/England/ALDP-A50CA9/2020\|EPI_ISL_597657\|20 | ···················· | ···················· | ···················· | ···················· | ···················· | ···················· |
| N | 7971 | hCoV-19/England/ALDP-A4FA49/2020\|EPI_ISL_597659\|20 | ···················· | ···················· | ···················· | ···················· | ···················· | ···················· |

| | | | | | | | | |
|---|---|---|---|---|---|---|---|---|
| N | 7972 | hCoV-19/England/ALDP-A50526/2020\|EPI_ISL_597660\|20 | ···················· | ···················· | ····················· | ···················· | ······T············· | ···················· |
| N | 7973 | hCoV-19/England/ALDP-A4F9E2/2020\|EPI_ISL_597661\|20 | ···················· | ···················· | ····················· | ···················· | ···················· | ···················· |
| N | 7974 | hCoV-19/England/ALDP-A4FB73/2020\|EPI_ISL_597662\|20 | ···················· | ···················· | ····················· | ···················· | ···················· | ···················· |
| N | 7975 | hCoV-19/England/ALDP-A4FC61/2020\|EPI_ISL_597664\|20 | ···················· | ···················· | ····················· | ···················· | ···················· | ···················· |
| N | 7976 | hCoV-19/England/ALDP-A50FFB/2020\|EPI_ISL_597665\|20 | ···················· | ···················· | ····················· | ···················· | ···················· | ···················· |
| N | 7977 | hCoV-19/England/ALDP-A50474/2020\|EPI_ISL_597666\|20 | ···················· | ···················· | ····················· | ···················· | ···················· | ···················· |
| N | 7978 | hCoV-19/England/ALDP-A50957/2020\|EPI_ISL_597667\|20 | ···················· | ·A·················· | ····················· | ···················· | ···················· | ···················· |
| N | 7979 | hCoV-19/England/ALDP-A5083C/2020\|EPI_ISL_597668\|20 | ···················· | ···················· | ····················· | ···················· | ···················· | ···················· |
| N | 7980 | hCoV-19/England/ALDP-A50E67/2020\|EPI_ISL_597669\|20 | ···················· | ···················· | ····················· | ···················· | ···················· | ···················· |
| N | 7981 | hCoV-19/England/ALDP-A507F3/2020\|EPI_ISL_597670\|20 | ···················· | ···················· | ····················· | ···················· | ···················· | ···················· |
| N | 7982 | hCoV-19/England/ALDP-A50BE8/2020\|EPI_ISL_597671\|20 | ···················· | ···················· | ····················· | ···················· | T··················· | ···················· |
| N | 7983 | hCoV-19/England/ALDP-A4FE10/2020\|EPI_ISL_597672\|20 | ···················· | ···················· | ····················· | ···················· | ···················· | ···················· |
| N | 7984 | hCoV-19/England/ALDP-A50CB8/2020\|EPI_ISL_597673\|20 | ···················· | ···················· | ····················· | ···················· | ···················· | ···················· |
| N | 7985 | hCoV-19/England/ALDP-A50DB5/2020\|EPI_ISL_597674\|20 | ···················· | ···················· | ····················· | ···················· | ···················· | ···················· |
| N | 7986 | hCoV-19/England/ALDP-A4FAD0/2020\|EPI_ISL_597675\|20 | ···················· | ···················· | ····················· | ···················· | ···················· | ···················· |
| N | 7987 | hCoV-19/Scotland/QEUH-A203F8/2020\|EPI_ISL_598920\|2 | ···················· | ···················· | ····················· | ···················· | ···················· | ···················· |
| N | 7988 | hCoV-19/Scotland/QEUH-A1FE46/2020\|EPI_ISL_598921\|2 | ···················· | ···················· | ····················· | ···················· | ···················· | ···················· |
| N | 7989 | hCoV-19/Scotland/QEUH-A225A5/2020\|EPI_ISL_598922\|2 | ···················· | ···················· | ····················· | ···················· | ···················· | ···················· |
| N | 7990 | hCoV-19/Scotland/QEUH-A1F6E4/2020\|EPI_ISL_598923\|2 | ···················· | ···················· | ····················· | ···················· | ···················· | ···················· |
| N | 7991 | hCoV-19/Scotland/QEUH-A20E06/2020\|EPI_ISL_598924\|2 | ···················· | ···················· | ····················· | ···················· | ···················· | ···················· |
| N | 7992 | hCoV-19/Scotland/QEUH-A1FF25/2020\|EPI_ISL_598925\|2 | ···················· | ···················· | ····················· | ···················· | ···················· | ···················· |
| N | 7993 | hCoV-19/Scotland/QEUH-A1FA51/2020\|EPI_ISL_598926\|2 | ···················· | ···················· | ····················· | ···················· | ···················· | ···················· |
| N | 7994 | hCoV-19/Scotland/QEUH-A1FEFB/2020\|EPI_ISL_598929\|2 | ···················· | ···················· | ····················· | ···················· | ···················· | ···················· |
| N | 7995 | hCoV-19/Scotland/QEUH-A22718/2020\|EPI_ISL_598930\|2 | ···················· | ···················· | ····················· | ···················· | ···················· | ···················· |
| N | 7996 | hCoV-19/Scotland/QEUH-A1F769/2020\|EPI_ISL_598931\|2 | ···················· | ···················· | ····················· | ···················· | ···················· | ···················· |
| N | 7997 | hCoV-19/Scotland/QEUH-A1FEEC/2020\|EPI_ISL_598933\|2 | ···················· | ···················· | ····················· | ···················· | ···················· | ···················· |
| N | 7998 | hCoV-19/Scotland/QEUH-A20DEB/2020\|EPI_ISL_598934\|2 | ···················· | ···················· | ····················· | ···················· | ···················· | ···················· |
| N | 7999 | hCoV-19/Scotland/QEUH-A210B4/2020\|EPI_ISL_598935\|2 | ···················· | ···················· | ····················· | ···················· | ···················· | ···················· |
| N | 8000 | hCoV-19/Scotland/QEUH-A1FA9D/2020\|EPI_ISL_598937\|2 | ···················· | ···················· | ····················· | ···················· | ···················· | ···················· |
| N | 8001 | hCoV-19/Scotland/QEUH-A20D81/2020\|EPI_ISL_598938\|2 | ···················· | ···················· | ····················· | ···················· | ···················· | ···················· |
| N | 8002 | hCoV-19/Scotland/QEUH-A20E7F/2020\|EPI_ISL_598939\|2 | ···················· | ···················· | ····················· | ···················· | ···················· | ···················· |
| N | 8003 | hCoV-19/Scotland/QEUH-A1FD3A/2020\|EPI_ISL_598940\|2 | ···················· | ···················· | ····················· | ···················· | ···················· | ···················· |
| N | 8004 | hCoV-19/Scotland/QEUH-A1F505/2020\|EPI_ISL_598963\|2 | ···················· | ···················· | ····················· | ···················· | ···················· | ···················· |
| N | 8005 | hCoV-19/Scotland/QEUH-A1F5AB/2020\|EPI_ISL_598964\|2 | ···················· | ···················· | ····················· | ···················· | ···················· | ···················· |
| N | 8006 | hCoV-19/Scotland/QEUH-A1FB30/2020\|EPI_ISL_598965\|2 | ···················· | ···················· | ····················· | ···················· | ···················· | ···················· |
| N | 8007 | hCoV-19/Scotland/QEUH-A1F7E1/2020\|EPI_ISL_598966\|2 | ···················· | ···················· | ····················· | ···················· | ···················· | ···················· |
| N | 8008 | hCoV-19/Scotland/QEUH-A1F57E/2020\|EPI_ISL_598967\|2 | ···················· | ···················· | ····················· | ···················· | ···················· | ···················· |
| N | 8009 | hCoV-19/Scotland/QEUH-A210C3/2020\|EPI_ISL_598968\|2 | ···················· | ···················· | ····················· | ···················· | ···················· | ···················· |
| N | 8010 | hCoV-19/Scotland/QEUH-A1F514/2020\|EPI_ISL_598969\|2 | ···················· | ···················· | ····················· | ···················· | ···················· | ···················· |
| N | 8011 | hCoV-19/Scotland/QEUH-A1FE64/2020\|EPI_ISL_598971\|2 | ···················· | ···················· | ····················· | ···················· | ···················· | ···················· |
| N | 8012 | hCoV-19/Scotland/QEUH-A1F972/2020\|EPI_ISL_598972\|2 | ···················· | ···················· | ····················· | ···················· | ···················· | ···················· |
| N | 8013 | hCoV-19/Scotland/QEUH-A1F64E/2020\|EPI_ISL_598973\|2 | ···················· | ···················· | ····················· | ···················· | ···················· | ···················· |
| N | 8014 | hCoV-19/Scotland/QEUH-A1FD94/2020\|EPI_ISL_598974\|2 | ···················· | ···················· | ····················· | ···················· | ···················· | ···················· |
| N | 8015 | hCoV-19/Scotland/QEUH-A1FD58/2020\|EPI_ISL_598975\|2 | ···················· | ···················· | ····················· | ···················· | ···················· | ···················· |
| N | 8016 | hCoV-19/Scotland/QEUH-A20DFA/2020\|EPI_ISL_598976\|2 | ···················· | ···················· | ···········Y········· | ···················· | ···················· | ···················· |
| N | 8017 | hCoV-19/Scotland/QEUH-A20923/2020\|EPI_ISL_598977\|2 | ···················· | ···················· | ····················· | ···················· | ···················· | ···················· |
| N | 8018 | hCoV-19/Scotland/QEUH-A2250F/2020\|EPI_ISL_598978\|2 | ···················· | ···················· | ····················· | ···················· | ···················· | ···················· |
| N | 8019 | hCoV-19/Scotland/QEUH-A1FB7C/2020\|EPI_ISL_598979\|2 | ···················· | ···················· | ····················· | ···················· | ···················· | ···················· |
| N | 8020 | hCoV-19/Scotland/QEUH-A1FFCB/2020\|EPI_ISL_598980\|2 | ···················· | ···················· | ····················· | ···················· | ···················· | ···················· |
| N | 8021 | hCoV-19/Scotland/QEUH-A20431/2020\|EPI_ISL_598981\|2 | ···················· | ···················· | ····················· | ···················· | ···················· | ···················· |
| N | 8022 | hCoV-19/Scotland/QEUH-A1FD2B/2020\|EPI_ISL_598982\|2 | ···················· | ···················· | ····················· | ···················· | ···················· | ···················· |
| N | 8023 | hCoV-19/Scotland/QEUH-A1F963/2020\|EPI_ISL_598983\|2 | ···················· | ···················· | ····················· | ···················· | ···················· | ···················· |
| N | 8024 | hCoV-19/Scotland/QEUH-A1FC3D/2020\|EPI_ISL_598985\|2 | ···················· | ···················· | ····················· | ···················· | ···················· | ···················· |
| N | 8025 | hCoV-19/Scotland/QEUH-A20E33/2020\|EPI_ISL_598986\|2 | ···················· | ···················· | ····················· | ···················· | ···················· | ···················· |
| N | 8026 | hCoV-19/Scotland/QEUH-A1FB12/2020\|EPI_ISL_598987\|2 | ···················· | ···················· | ····················· | ···················· | ···················· | ···················· |
| N | 8027 | hCoV-19/Scotland/QEUH-A1F5BA/2020\|EPI_ISL_598988\|2 | ···················· | ···················· | ····················· | ···················· | ···················· | ···················· |
| N | 8028 | hCoV-19/Scotland/QEUH-A1FA60/2020\|EPI_ISL_598989\|2 | ···················· | ···················· | ····················· | ···················· | ···················· | ···················· |
| N | 8029 | hCoV-19/Scotland/QEUH-A1FBB8/2020\|EPI_ISL_598991\|2 | ···················· | ···················· | ····················· | ···················· | ···················· | ···················· |
| N | 8030 | hCoV-19/Scotland/QEUH-A2097E/2020\|EPI_ISL_598992\|2 | ···················· | ···················· | ····················· | ···················· | ···················· | ···················· |
| N | 8031 | hCoV-19/Scotland/QEUH-A1FA42/2020\|EPI_ISL_598993\|2 | ···················· | ···················· | ····················· | ···················· | ···················· | ···················· |
| N | 8032 | hCoV-19/Scotland/QEUH-A1F8EE/2020\|EPI_ISL_598994\|2 | ···················· | ···················· | ····················· | ···················· | ···················· | ···················· |
| N | 8033 | hCoV-19/Scotland/QEUH-A1FC1F/2020\|EPI_ISL_598995\|2 | ···················· | ···················· | ····················· | ···················· | ···················· | ···················· |
| N | 8034 | hCoV-19/Scotland/QEUH-A1F936/2020\|EPI_ISL_598996\|2 | ···················· | ···················· | ····················· | ···················· | ···················· | ···················· |
| N | 8035 | hCoV-19/Scotland/QEUH-A1F5F6/2020\|EPI_ISL_598997\|2 | ···················· | ···················· | ····················· | ···················· | ···················· | ···················· |
| N | 8036 | hCoV-19/Scotland/QEUH-A1FF16/2020\|EPI_ISL_598998\|2 | ···················· | ···················· | ····················· | ···················· | ···················· | ···················· |
| N | 8037 | hCoV-19/Scotland/QEUH-A225E1/2020\|EPI_ISL_598999\|2 | ···················· | ···················· | ····················· | ···················· | ···················· | ···················· |
| N | 8038 | hCoV-19/Wales/MILK-9E0AF0/2020\|EPI_ISL_601310\|2020 | ···················· | ···················· | ····················· | ···················· | ···················· | ···················· |
| N | 8039 | hCoV-19/England/MILK-9E0B48/2020\|EPI_ISL_601313\|20 | ···················· | ···················· | ····················· | ···················· | ···················· | ···················· |
| N | 8040 | hCoV-19/England/QEUH-9DAA53/2020\|EPI_ISL_601315\|20 | ···················· | ···················· | ····················· | ···················· | ···················· | ···················· |
| N | 8041 | hCoV-19/Wales/MILK-9E0C27/2020\|EPI_ISL_601316\|2020 | ···················· | ···················· | ····················· | ···················· | ···················· | ···················· |
| N | 8042 | hCoV-19/England/MILK-9E0BA2/2020\|EPI_ISL_601317\|20 | ···················· | ···················· | ····················· | ···················· | ···················· | ···················· |
| N | 8043 | hCoV-19/England/QEUH-9DA4BF/2020\|EPI_ISL_601319\|20 | ···················· | ···················· | ····················· | ···················· | ···················· | ···················· |
| N | 8044 | hCoV-19/England/MILK-9E096C/2020\|EPI_ISL_601320\|20 | ···················· | ···················· | ····················· | ···················· | ···················· | ···················· |
| N | 8045 | hCoV-19/Wales/MILK-9E051D/2020\|EPI_ISL_601321\|2020 | ···················· | ···················· | ····················· | ···················· | ···················· | ···················· |
| N | 8046 | hCoV-19/Wales/MILK-9E04C5/2020\|EPI_ISL_601322\|2020 | ···················· | ···················· | ····················· | ···················· | ···················· | ···················· |
| N | 8047 | hCoV-19/England/QEUH-9DC268/2020\|EPI_ISL_601323\|20 | ···················· | ···················· | ····················· | ···················· | ···················· | ···················· |
| N | 8048 | hCoV-19/England/MILK-9E0753/2020\|EPI_ISL_601324\|20 | ···················· | ·A·················· | ····················· | ···················· | ···················· | ··A················· |
| N | 8049 | hCoV-19/England/MILK-9E0C81/2020\|EPI_ISL_601325\|20 | ···················· | ···················· | ····················· | ···················· | ···················· | ···················· |
| N | 8050 | hCoV-19/England/MILK-9E0629/2020\|EPI_ISL_601326\|20 | ···················· | ···················· | ····················· | ···················· | ···················· | ···················· |
| N | 8051 | hCoV-19/England/MILK-9E0717/2020\|EPI_ISL_601327\|20 | ···················· | ···················· | ····················· | ···················· | ···················· | ···················· |

```
N 8052 hCoV-19/South Africa/KRISP-K002819/2020|EPI_ISL_60 ···················· ···················· ···················· ···················· ···················· ····················
N 8053 hCoV-19/South Africa/KRISP-K002820/2020|EPI_ISL_60 ···················· ···················· ···················· ···················· ···················· ····················
N 8054 hCoV-19/South Africa/KRISP-K002821/2020|EPI_ISL_60 ···················· ···················· ···················· ···················· ···················· ····················
N 8055 hCoV-19/South Africa/KRISP-K002826/2020|EPI_ISL_60 ···················· ···················· ···················· ···················· ···················· ····················
N 8056 hCoV-19/South Africa/KRISP-K002827/2020|EPI_ISL_60 ···················· ···················· ···················· ···················· ···················· ····················
N 8057 hCoV-19/South Africa/KRISP-K002828/2020|EPI_ISL_60 ···················· ···················· ···················· ···················· ···T················ ····················
N 8058 hCoV-19/South Africa/KRISP-K002829/2020|EPI_ISL_60 ···················· ···················· ···················· ···················· ···················· ····················
N 8059 hCoV-19/South Africa/KRISP-K002830/2020|EPI_ISL_60 ···················· ···················· ···················· ···················· ···················· ····················
N 8060 hCoV-19/South Africa/KRISP-K002831/2020|EPI_ISL_60 ···················· ···················· ···················· ···················· ···················· ····················
N 8061 hCoV-19/South Africa/KRISP-K002833/2020|EPI_ISL_60 ···················· ·A·················· ···················· ···················· ···················· ····················
N 8062 hCoV-19/South Africa/KRISP-K002836/2020|EPI_ISL_60 ···················· ···················· ···················· ···················· ···················· ····················
N 8063 hCoV-19/South Africa/KRISP-K002837/2020|EPI_ISL_60 ···················· ···················· ···················· ···················· ···················· ····················
N 8064 hCoV-19/South Africa/KRISP-K002840/2020|EPI_ISL_60 ···················· ···················· ···················· ···················· ···················· ····················
N 8065 hCoV-19/South Africa/KRISP-K002841/2020|EPI_ISL_60 ···················· ···················· ···················· ···················· ···················· ····················
N 8066 hCoV-19/South Africa/KRISP-K002842/2020|EPI_ISL_60 ···················· ···················· ···················· ···················· ···················· ····················
N 8067 hCoV-19/USA/CA-QDX-1607/2020|EPI_ISL_603902|2020-0 ···················· ···················· ···················· ···················· ···················· ····················
N 8068 hCoV-19/USA/CA-QDX-1609/2020|EPI_ISL_603903|2020-0 ···················· ···················· ···················· ···················· ···················· ····················
N 8069 hCoV-19/USA/CA-QDX-1622/2020|EPI_ISL_603904|2020-0 ···················· ···················· ···················· ···················· ···················· ····················
N 8070 hCoV-19/USA/CA-QDX-1623/2020|EPI_ISL_603905|2020-0 ···················· ···················· ···················· ···················· ···················· ····················
N 8071 hCoV-19/USA/CA-QDX-1624/2020|EPI_ISL_603906|2020-0 ···················· ···················· ···················· ···················· ···················· ····················
N 8072 hCoV-19/USA/CA-QDX-1625/2020|EPI_ISL_603907|2020-0 ···················· ···················· ···················· ···················· ···················· ····················
N 8073 hCoV-19/USA/CA-QDX-1629/2020|EPI_ISL_603908|2020-0 ···················· ···················· ···················· ···················· ···················· ····················
N 8074 hCoV-19/USA/CA-QDX-1633/2020|EPI_ISL_603909|2020-0 ···················· ···················· ···················· ···················· ···················· ····················
N 8075 hCoV-19/USA/CA-QDX-1640/2020|EPI_ISL_603910|2020-0 ···················· ···················· ···················· ···················· ···················· ····················
N 8076 hCoV-19/USA/CA-QDX-1641/2020|EPI_ISL_603911|2020-0 ···················· ···················· ···················· ···················· ···················· ····················
N 8077 hCoV-19/USA/CA-QDX-1644/2020|EPI_ISL_603912|2020-0 ···················· ···················· ···················· ···················· ···················· ····················
N 8078 hCoV-19/USA/CA-QDX-1651/2020|EPI_ISL_603913|2020-0 ···················· ···················· ···················· ···················· ···················· ····················
N 8079 hCoV-19/USA/CA-QDX-1656/2020|EPI_ISL_603915|2020-0 ···················· ···················· ···················· ···················· ···················· ····················
N 8080 hCoV-19/USA/CA-QDX-1657/2020|EPI_ISL_603916|2020-0 ···················· ···················· ···················· ···················· ···················· ····················
N 8081 hCoV-19/USA/CA-QDX-1661/2020|EPI_ISL_603917|2020-0 ···················· ···················· ···················· ···················· ···················· ····················
N 8082 hCoV-19/USA/CA-QDX-1662/2020|EPI_ISL_603918|2020-0 ···················· ···················· ···················· ···················· ···················· ····················
N 8083 hCoV-19/USA/CA-QDX-1668/2020|EPI_ISL_603919|2020-0 ···················· ···················· ···················· ···················· ···················· ····················
N 8084 hCoV-19/USA/CA-QDX-1669/2020|EPI_ISL_603920|2020-0 ···················· ···················· ···················· ···················· ···················· ····················
N 8085 hCoV-19/USA/CA-QDX-1670/2020|EPI_ISL_603921|2020-0 ···················· ···················· ···················· ···················· ···················· ····················
N 8086 hCoV-19/USA/CA-QDX-1671/2020|EPI_ISL_603922|2020-0 ···················· ···················· ···················· ···················· ···················· ····················
N 8087 hCoV-19/USA/CA-QDX-1674/2020|EPI_ISL_603923|2020-0 ···················· ···················· ···················· ···················· ···················· ····················
N 8088 hCoV-19/USA/CA-QDX-1675/2020|EPI_ISL_603924|2020-0 ···················· ···················· ···················· ···················· ···················· ····················
N 8089 hCoV-19/USA/CA-QDX-1676/2020|EPI_ISL_603925|2020-0 ···················· ···················· ···················· ···················· ···················· ····················
N 8090 hCoV-19/USA/CA-QDX-1677/2020|EPI_ISL_603926|2020-0 ···················· ···················· ···················· ···················· ···················· ····················
N 8091 hCoV-19/USA/CA-QDX-1679/2020|EPI_ISL_603927|2020-0 ···················· ···················· ···················· ···················· ···················· ····················
N 8092 hCoV-19/USA/CA-QDX-1680/2020|EPI_ISL_603928|2020-0 ···················· ···················· ···················· ···················· ···················· ····················
N 8093 hCoV-19/USA/CA-QDX-1682/2020|EPI_ISL_603929|2020-0 ···················· ···················· ···················· ···················· ···················· ····················
N 8094 hCoV-19/USA/CA-QDX-1684/2020|EPI_ISL_603930|2020-0 ···················· ···················· ···················· ···················· ···················· ····················
N 8095 hCoV-19/USA/CA-QDX-1685/2020|EPI_ISL_603931|2020-0 ···················· ···················· ···················· ···················· ···················· ····················
N 8096 hCoV-19/USA/CA-QDX-1689/2020|EPI_ISL_603932|2020-0 ···················· ···················· ···················· ···················· ···················· ····················
N 8097 hCoV-19/USA/CA-QDX-1693/2020|EPI_ISL_603933|2020-0 ···················· ···················· ···················· ···················· ···················· ····················
N 8098 hCoV-19/USA/CA-QDX-1700/2020|EPI_ISL_603934|2020-0 ···················· ···················· ···················· ···················· ···················· ····················
N 8099 hCoV-19/USA/FL-QDX-1590/2020|EPI_ISL_603935|2020-0 ···················· ···················· ···················· ···················· ···················· ····················
N 8100 hCoV-19/USA/FL-QDX-1606/2020|EPI_ISL_603936|2020-0 ···················· ···················· ···················· ···················· ···················· ····················
N 8101 hCoV-19/USA/FL-QDX-1616/2020|EPI_ISL_603937|2020-0 ···················· ···················· ···················· ···················· ···················· ····················
N 8102 hCoV-19/USA/FL-QDX-1620/2020|EPI_ISL_603938|2020-0 ···················· ···················· ···················· ···················· ···················· ····················
N 8103 hCoV-19/USA/FL-QDX-1628/2020|EPI_ISL_603939|2020-0 ···················· ···················· ···················· ···················· ···················· ····················
N 8104 hCoV-19/USA/FL-QDX-1638/2020|EPI_ISL_603940|2020-0 ···················· ···················· ···················· ···················· ···················· ····················
N 8105 hCoV-19/USA/FL-QDX-1658/2020|EPI_ISL_603941|2020-0 ···················· ···················· ···················· ···················· ···················· ····················
N 8106 hCoV-19/USA/IN-QDX-1569/2020|EPI_ISL_603942|2020-0 ···················· ···················· ···················· ···················· ···················· ····················
N 8107 hCoV-19/USA/MA-QDX-1696/2020|EPI_ISL_603944|2020-0 ···················· ···················· ···················· ···················· ···················· ····················
N 8108 hCoV-19/USA/MI-QDX-1563/2020|EPI_ISL_603945|2020-0 ···················· ···················· ···················· ···················· ···················· ····················
N 8109 hCoV-19/USA/MI-QDX-1683/2020|EPI_ISL_603970|2020-0 ···················· ···················· ···················· ···················· ···················· ····················
N 8110 hCoV-19/USA/MI-QDX-1686/2020|EPI_ISL_603971|2020-0 ···················· ···················· ···················· ···················· ···················· ····················
N 8111 hCoV-19/USA/MI-QDX-1687/2020|EPI_ISL_603972|2020-0 ···················· ···················· ···················· ···················· ···················· ····················
N 8112 hCoV-19/USA/MI-QDX-1690/2020|EPI_ISL_603973|2020-0 ···················· ···················· ···················· ···················· ···················· ····················
N 8113 hCoV-19/USA/MN-QDX-1561/2020|EPI_ISL_603974|2020-0 ···················· ···················· ···················· ···················· ···················· ····················
N 8114 hCoV-19/USA/MN-QDX-1582/2020|EPI_ISL_603978|2020-0 ···················· ···················· ···················· ···················· ···················· ····················
N 8115 hCoV-19/USA/MN-QDX-1593/2020|EPI_ISL_603979|2020-0 ···················· ···················· ···················· ···················· ···················· ····················
N 8116 hCoV-19/USA/MN-QDX-1597/2020|EPI_ISL_603980|2020-0 ···················· ···················· ···················· ···················· ···················· ····················
N 8117 hCoV-19/USA/MN-QDX-1601/2020|EPI_ISL_603981|2020-0 ···················· ···················· ···················· ···················· ···················· ····················
N 8118 hCoV-19/USA/MN-QDX-1602/2020|EPI_ISL_603982|2020-0 ···················· ···················· ···················· ···················· ···················· ····················
N 8119 hCoV-19/USA/MN-QDX-1603/2020|EPI_ISL_603983|2020-0 ···················· ···················· ···················· ···················· ···················· ····················
N 8120 hCoV-19/USA/MN-QDX-1604/2020|EPI_ISL_603984|2020-0 ···················· ···················· ···················· ···················· ···················· ····················
N 8121 hCoV-19/USA/MN-QDX-1608/2020|EPI_ISL_603985|2020-0 ···················· ···················· ···················· ···················· ···················· ····················
N 8122 hCoV-19/USA/MN-QDX-1611/2020|EPI_ISL_603986|2020-0 ···················· ···················· ···················· ···················· ···················· ····················
N 8123 hCoV-19/USA/MN-QDX-1615/2020|EPI_ISL_603987|2020-0 ···················· ···················· ···················· ···················· ···················· ····················
N 8124 hCoV-19/USA/MN-QDX-1617/2020|EPI_ISL_603988|2020-0 ···················· ···················· ···················· ···················· ···················· ····················
N 8125 hCoV-19/USA/MN-QDX-1618/2020|EPI_ISL_603989|2020-0 ···················· ···················· ···················· ···················· ···················· ····················
O    1 hCoV-19/England/MILK-A7D0AA/2020|EPI_ISL_606900|20 ···················· ···················· ···················· ···················· ···················· ····················
O    2 hCoV-19/England/MILK-A7D0E6/2020|EPI_ISL_606902|20 ···················· ···················· ···················· ···················· ···················· ····················
O    3 hCoV-19/England/MILK-A7976D/2020|EPI_ISL_606903|20 ···················· ···················· ···················· ···················· ···················· ····················
O    4 hCoV-19/England/MILK-A7A535/2020|EPI_ISL_606906|20 ···················· ···················· ···················· ···················· ···················· ····················
O    5 hCoV-19/England/MILK-A7A553/2020|EPI_ISL_606907|20 ···················· ···················· ···················· ···················· ···················· ····················
O    6 hCoV-19/England/MILK-A7A210/2020|EPI_ISL_606908|20 ···················· ···················· ···················· ···················· ···················· ····················
```

| | | | | | | | |
|---|---|---|---|---|---|---|---|
| O | 7 hCoV-19/England/MILK-A79633/2020\|EPI_ISL_606910\|20 | .................... | .................... | .................... | .................... | .................... | .................... |
| O | 8 hCoV-19/England/MILK-A79967/2020\|EPI_ISL_606911\|20 | .................... | .................... | .................... | .................... | .................... | .................... |
| O | 9 hCoV-19/England/MILK-A7D06E/2020\|EPI_ISL_606912\|20 | .................... | .................... | .................... | .................... | .................... | .................... |
| O | 10 hCoV-19/England/MILK-A7CCB6/2020\|EPI_ISL_606913\|20 | .................... | .................... | .................... | .................... | .....T.............. | .................... |
| O | 11 hCoV-19/England/MILK-A79B16/2020\|EPI_ISL_606914\|20 | .................... | .................... | .................... | .................... | .................... | .................... |
| O | 12 hCoV-19/England/MILK-A7CE56/2020\|EPI_ISL_606915\|20 | .................... | .................... | .................... | .................... | .................... | .................... |
| O | 13 hCoV-19/England/MILK-A79CB9/2020\|EPI_ISL_606917\|20 | .................... | .................... | .................... | .................... | .................... | .................... |
| O | 14 hCoV-19/England/MILK-A7CCD4/2020\|EPI_ISL_606918\|20 | .................... | .A.................. | .................... | .................... | .................... | ..A................. |
| O | 15 hCoV-19/England/MILK-A7AB8E/2020\|EPI_ISL_606919\|20 | .................... | .................... | .................... | .................... | .................... | .................... |
| O | 16 hCoV-19/England/MILK-A7CE83/2020\|EPI_ISL_606920\|20 | .................... | .................... | .................... | .................... | .................... | .................... |
| O | 17 hCoV-19/England/MILK-A7A650/2020\|EPI_ISL_606921\|20 | .................... | .................... | .................... | .................... | .....T.............. | .................... |
| O | 18 hCoV-19/England/MILK-A79D6B/2020\|EPI_ISL_606923\|20 | .................... | .................... | .................... | .................... | .................... | .................... |
| O | 19 hCoV-19/England/ALDP-A6FD4A/2020\|EPI_ISL_606925\|20 | .................... | .................... | .................... | .................... | .................... | .................... |
| O | 20 hCoV-19/England/ALDP-A6F32A/2020\|EPI_ISL_606926\|20 | .................... | .................... | .................... | .................... | .................... | .................... |
| O | 21 hCoV-19/England/ALDP-A6FE0B/2020\|EPI_ISL_606927\|20 | .................... | .................... | .................... | .................... | .................... | .................... |
| O | 22 hCoV-19/England/ALDP-A6DCF4/2020\|EPI_ISL_606928\|20 | .................... | .................... | .................... | .................... | .................... | .................... |
| O | 23 hCoV-19/England/ALDP-A6C93A/2020\|EPI_ISL_606929\|20 | .................... | .A.................. | .................... | .................... | .................... | ..A................. |
| O | 24 hCoV-19/England/ALDP-A6CA0A/2020\|EPI_ISL_606930\|20 | .................... | .................... | .................... | .................... | .................... | .................... |
| O | 25 hCoV-19/England/ALDP-A6DD79/2020\|EPI_ISL_606931\|20 | .................... | .................... | .................... | .................... | .................... | .................... |
| O | 26 hCoV-19/England/ALDP-A6D5AE/2020\|EPI_ISL_606932\|20 | .................... | .................... | .................... | .................... | .................... | .................... |
| O | 27 hCoV-19/England/ALDP-A6F366/2020\|EPI_ISL_606933\|20 | .................... | .................... | .................... | .................... | .................... | .................... |
| O | 28 hCoV-19/England/ALDP-A6E130/2020\|EPI_ISL_606934\|20 | .................... | .................... | .................... | .................... | .................... | .................... |
| O | 29 hCoV-19/England/ALDP-A6CAA0/2020\|EPI_ISL_606936\|20 | .................... | .................... | .................... | .................... | .................... | .................... |
| O | 30 hCoV-19/England/ALDP-A6EDD2/2020\|EPI_ISL_606937\|20 | .................... | .A.................. | .................... | .................... | .................... | .................... |
| O | 31 hCoV-19/England/ALDP-A6ED96/2020\|EPI_ISL_606938\|20 | .................... | .................... | .................... | .................... | .................... | .................... |
| O | 32 hCoV-19/England/ALDP-A6D9DF/2020\|EPI_ISL_606939\|20 | .................... | .................... | .................... | .................... | .................... | .................... |
| O | 33 hCoV-19/England/ALDP-A6E103/2020\|EPI_ISL_606940\|20 | .................... | .A.................. | .................... | .................... | .................... | ..A................. |
| O | 34 hCoV-19/England/ALDP-A6F7B5/2020\|EPI_ISL_606941\|20 | .................... | .................... | .................... | .................... | .................... | .................... |
| O | 35 hCoV-19/England/ALDP-A6F393/2020\|EPI_ISL_606942\|20 | .................... | .................... | .................... | .................... | .................... | .................... |
| O | 36 hCoV-19/England/ALDP-A6F76A/2020\|EPI_ISL_606943\|20 | .................... | .................... | .................... | .................... | .................... | .................... |
| O | 37 hCoV-19/England/ALDP-A6F296/2020\|EPI_ISL_606944\|20 | .................... | .................... | .................... | .................... | .................... | .................... |
| O | 38 hCoV-19/England/ALDP-A6FACB/2020\|EPI_ISL_606945\|20 | .................... | .................... | .................... | .................... | .................... | .................... |
| O | 39 hCoV-19/England/ALDP-A6F2F0/2020\|EPI_ISL_606946\|20 | .................... | .................... | .................... | .................... | .................... | .................... |
| O | 40 hCoV-19/England/ALDP-A6FBB9/2020\|EPI_ISL_606947\|20 | .................... | .................... | .................... | .................... | .................... | .................... |
| O | 41 hCoV-19/England/ALDP-A6F788/2020\|EPI_ISL_606949\|20 | .................... | .................... | .................... | .................... | .................... | .................... |
| O | 42 hCoV-19/England/ALDP-A6CBDA/2020\|EPI_ISL_606950\|20 | .................... | .A.................. | .................... | .................... | .................... | ..A................. |
| O | 43 hCoV-19/England/ALDP-A6FBD7/2020\|EPI_ISL_606951\|20 | .................... | .................... | .................... | .................... | .................... | .................... |
| O | 44 hCoV-19/England/ALDP-A6EADB/2020\|EPI_ISL_606952\|20 | .................... | .................... | .................... | .................... | .................... | .................... |
| O | 45 hCoV-19/England/ALDP-A6CA55/2020\|EPI_ISL_606953\|20 | .................... | .................... | .................... | .................... | .................... | .................... |
| O | 46 hCoV-19/England/ALDP-A6EEFD/2020\|EPI_ISL_606954\|20 | .................... | .................... | .................... | .................... | .................... | .................... |
| O | 47 hCoV-19/England/ALDP-A6EBBA/2020\|EPI_ISL_606955\|20 | .................... | .................... | .................... | .................... | .................... | .................... |
| O | 48 hCoV-19/England/ALDP-A6E5AD/2020\|EPI_ISL_606956\|20 | .................... | .................... | .................... | .................... | .................... | .................... |
| O | 49 hCoV-19/England/ALDP-A6EC5D/2020\|EPI_ISL_606957\|20 | .................... | .................... | .................... | .................... | .................... | .................... |
| O | 50 hCoV-19/England/ALDP-A6EB23/2020\|EPI_ISL_606959\|20 | .................... | .................... | .................... | .................... | .................... | .................... |
| O | 51 hCoV-19/England/ALDP-A6E4DD/2020\|EPI_ISL_606961\|20 | .................... | .................... | .................... | .................... | .................... | .................... |
| O | 52 hCoV-19/England/ALDP-A6E640/2020\|EPI_ISL_606962\|20 | .................... | .................... | .................... | .................... | .................... | .................... |
| O | 53 hCoV-19/England/ALDP-A6DD5B/2020\|EPI_ISL_606963\|20 | .................... | .................... | .................... | .................... | .................... | .................... |
| O | 54 hCoV-19/England/ALDP-A6E30D/2020\|EPI_ISL_606964\|20 | .................... | .................... | .................... | .................... | .................... | .................... |
| O | 55 hCoV-19/England/ALDP-A6E279/2020\|EPI_ISL_606965\|20 | .................... | .................... | .................... | .................... | .................... | ..A................. |
| O | 56 hCoV-19/England/ALDP-A6EA9F/2020\|EPI_ISL_606966\|20 | .................... | .................... | .................... | .................... | .................... | .................... |
| O | 57 hCoV-19/England/ALDP-A6D623/2020\|EPI_ISL_606967\|20 | .................... | .................... | .................... | .................... | .................... | .................... |
| O | 58 hCoV-19/England/ALDP-A6FB7D/2020\|EPI_ISL_606968\|20 | .................... | .................... | .................... | .................... | .................... | .................... |
| O | 59 hCoV-19/England/ALDP-A6F2D2/2020\|EPI_ISL_606969\|20 | .................... | .................... | .................... | .................... | .................... | .................... |
| O | 60 hCoV-19/England/ALDP-A6F14E/2020\|EPI_ISL_606970\|20 | .................... | .................... | .................... | .................... | .................... | .................... |
| O | 61 hCoV-19/England/ALDP-A6F18A/2020\|EPI_ISL_606971\|20 | .................... | .................... | .................... | .................... | .................... | .................... |
| O | 62 hCoV-19/England/ALDP-A6C9EF/2020\|EPI_ISL_606972\|20 | .................... | .................... | .................... | .................... | .................... | .................... |
| O | 63 hCoV-19/England/ALDP-A6FB9B/2020\|EPI_ISL_606973\|20 | .................... | .................... | .................... | .................... | .................... | .................... |
| O | 64 hCoV-19/England/ALDP-A6EDE1/2020\|EPI_ISL_606974\|20 | .................... | .................... | .................... | .................... | .................... | .................... |
| O | 65 hCoV-19/England/ALDP-A6D562/2020\|EPI_ISL_606975\|20 | .................... | .................... | .................... | .................... | .................... | .................... |
| O | 66 hCoV-19/England/ALDP-A6FB40/2020\|EPI_ISL_606976\|20 | .................... | .................... | .................... | .................... | .................... | .................... |
| O | 67 hCoV-19/England/ALDP-A6FCA7/2020\|EPI_ISL_606978\|20 | .................... | .................... | .................... | .................... | .................... | .................... |
| O | 68 hCoV-19/England/ALDP-A6F1B7/2020\|EPI_ISL_606979\|20 | .................... | .................... | .................... | .................... | .................... | .................... |
| O | 69 hCoV-19/England/ALDP-A6C9C1/2020\|EPI_ISL_606980\|20 | .................... | .................... | .................... | .................... | .................... | .................... |
| O | 70 hCoV-19/England/ALDP-A6C9A3/2020\|EPI_ISL_606981\|20 | .................... | .................... | .................... | .................... | .................... | .................... |
| O | 71 hCoV-19/England/ALDP-A6F22D/2020\|EPI_ISL_606982\|20 | .................... | .................... | .................... | .................... | .................... | .................... |
| O | 72 hCoV-19/England/ALDP-A6EE48/2020\|EPI_ISL_606983\|20 | .................... | .A.................. | .................... | .................... | .................... | .................... |
| O | 73 hCoV-19/England/ALDP-A6EC4E/2020\|EPI_ISL_606984\|20 | .................... | .................... | .................... | .................... | .................... | .................... |
| O | 74 hCoV-19/England/ALDP-A6EB50/2020\|EPI_ISL_606986\|20 | .................... | .................... | .................... | .................... | .................... | .................... |
| O | 75 hCoV-19/England/ALDP-A6F80D/2020\|EPI_ISL_606988\|20 | .................... | .................... | .................... | .................... | .................... | .................... |
| O | 76 hCoV-19/England/ALDP-A6C9FE/2020\|EPI_ISL_606989\|20 | .................... | .................... | .................... | .................... | .................... | .................... |
| O | 77 hCoV-19/England/ALDP-A6E22E/2020\|EPI_ISL_606990\|20 | .................... | .................... | .................... | .................... | .................... | .................... |
| O | 78 hCoV-19/England/ALDP-A6E622/2020\|EPI_ISL_606991\|20 | .................... | .................... | .................... | .................... | .................... | .................... |
| O | 79 hCoV-19/England/ALDP-A6E65F/2020\|EPI_ISL_606992\|20 | .................... | .................... | .................... | .................... | .................... | .................... |
| O | 80 hCoV-19/England/ALDP-A6DCA9/2020\|EPI_ISL_606993\|20 | .................... | .................... | .................... | .................... | .................... | .................... |
| O | 81 hCoV-19/England/ALDP-A6F7F1/2020\|EPI_ISL_606994\|20 | .................... | .................... | .................... | .................... | .................... | .................... |
| O | 82 hCoV-19/England/ALDP-A6E3A3/2020\|EPI_ISL_606995\|20 | .................... | .................... | .................... | .................... | .................... | .................... |
| O | 83 hCoV-19/England/ALDP-A6DD6A/2020\|EPI_ISL_606996\|20 | .................... | .................... | .................... | .................... | .................... | .................... |
| O | 84 hCoV-19/England/ALDP-A6DD2E/2020\|EPI_ISL_606997\|20 | .................... | .................... | .................... | .................... | .................... | .................... |
| O | 85 hCoV-19/England/ALDP-A6E6AA/2020\|EPI_ISL_606998\|20 | .................... | .................... | .................... | .................... | .................... | .................... |
| O | 86 hCoV-19/England/ALDP-A6FC10/2020\|EPI_ISL_606999\|20 | .................... | .................... | .................... | .................... | .................... | .................... |

```
O  87 hCoV-19/England/ALDP-A68A68/2020|EPI_ISL_609130|20  ···················· ···················· ···················· ···················· ···················· ····················
O  88 hCoV-19/England/CAMC-A649F6/2020|EPI_ISL_609131|20  ···················· ···················· ···················· ···················· ···················· ····················
O  89 hCoV-19/England/ALDP-A68057/2020|EPI_ISL_609132|20  ···················· ···················· ···················· ···················· ···················· ····················
O  90 hCoV-19/England/ALDP-A69074/2020|EPI_ISL_609133|20  ···················· ···················· ···················· ···················· ···················· ····················
O  91 hCoV-19/England/ALDP-A6929C/2020|EPI_ISL_609135|20  ···················· ·A·················· ···················· ···················· ···················· ···A················
O  92 hCoV-19/England/MILK-A6BF1B/2020|EPI_ISL_609136|20  ···················· ···················· ···················· ···················· ···················· ····················
O  93 hCoV-19/England/MILK-A6BD5D/2020|EPI_ISL_609137|20  ···················· ···················· ···················· ···················· ···················· ····················
O  94 hCoV-19/England/MILK-A6BD8A/2020|EPI_ISL_609138|20  ···················· ·A·················· ···················· ···················· ···················· ···A················
O  95 hCoV-19/England/MILK-A6BE5A/2020|EPI_ISL_609139|20  ···················· ···················· ···················· ···················· ···················· ····················
O  96 hCoV-19/England/ALDP-A67E8B/2020|EPI_ISL_609141|20  ···················· ·A·················· ···················· ···················· ···················· ····················
O  97 hCoV-19/England/CAMC-A64E9D/2020|EPI_ISL_609142|20  ···················· ···················· ···················· ···················· ···················· ····················
O  98 hCoV-19/England/CAMC-A64598/2020|EPI_ISL_609143|20  ···················· ···················· ···················· ···················· ···················· ····················
O  99 hCoV-19/England/ALDP-A69065/2020|EPI_ISL_609144|20  ···················· ···················· ···················· ···················· ···················· ····················
O 100 hCoV-19/England/ALDP-A680FD/2020|EPI_ISL_609146|20  ···················· ···················· ···················· ···················· ···················· ····················
O 101 hCoV-19/England/ALDP-A691F9/2020|EPI_ISL_609147|20  ···················· ···················· ···················· ···················· ···················· ····················
O 102 hCoV-19/England/ALDP-A67F88/2020|EPI_ISL_609148|20  ···················· ···················· ···················· ···················· ···················· ····················
O 103 hCoV-19/England/MILK-A6C0F9/2020|EPI_ISL_609149|20  ···················· ···················· ···················· ···················· ···················· ····················
O 104 hCoV-19/England/ALDP-A67FC4/2020|EPI_ISL_609150|20  ···················· ···················· ···················· ···················· ···················· ····················
O 105 hCoV-19/England/ALDP-A694D2/2020|EPI_ISL_609152|20  ···················· ···················· ···················· ···················· ···················· ····················
O 106 hCoV-19/England/ALDP-A692C9/2020|EPI_ISL_609153|20  ···················· ···················· ···················· ···················· ···················· ····················
O 107 hCoV-19/England/ALDP-A68AB3/2020|EPI_ISL_609154|20  ···················· ···················· ···················· ···················· ···················· ····················
O 108 hCoV-19/England/CAMC-A6461D/2020|EPI_ISL_609156|20  ···················· ···················· ···················· ···················· ···················· ····················
O 109 hCoV-19/England/ALDP-A68C17/2020|EPI_ISL_609157|20  ···················· ···················· ···················· ···················· ···················· ····················
O 110 hCoV-19/Wales/CAMC-A58A5C/2020|EPI_ISL_609159|2020  ···················· ···················· ···················· ···················· ···················· ····················
O 111 hCoV-19/Scotland/CAMC-A58CA1/2020|EPI_ISL_609171|2  ···················· ···················· ···················· ···················· ···················· ····················
O 112 hCoV-19/Scotland/CAMC-A586C1/2020|EPI_ISL_609172|2  ···················· ···················· ···················· ···················· ···················· ····················
O 113 hCoV-19/Wales/CAMC-A58DBD/2020|EPI_ISL_609173|2020  ···················· ···················· ···················· ···················· ···················· ····················
O 114 hCoV-19/England/CAMC-A59657/2020|EPI_ISL_609174|20  ···················· ···················· ···················· ···················· ···················· ····················
O 115 hCoV-19/Scotland/CAMC-A581B1/2020|EPI_ISL_609175|2  ···················· ···················· ···················· ···················· ···················· ····················
O 116 hCoV-19/Wales/CAMC-A588CB/2020|EPI_ISL_609176|2020  ···················· ···················· ···················· ···················· ···················· ····················
O 117 hCoV-19/Scotland/CAMC-A580D2/2020|EPI_ISL_609177|2  ···················· ···················· ···················· ···················· ···················· ····················
O 118 hCoV-19/Scotland/QEUH-A5E643/2020|EPI_ISL_609178|2  ···················· ···················· ···················· ···················· ···················· ····················
O 119 hCoV-19/Scotland/QEUH-A5DF76/2020|EPI_ISL_609179|2  ···················· ···················· ···················· ···················· ···················· ····················
O 120 hCoV-19/Wales/CAMC-A57501/2020|EPI_ISL_609180|2020  ···················· ···················· ···················· ···················· ···················· ····················
O 121 hCoV-19/Scotland/CAMC-A570B5/2020|EPI_ISL_609181|2  ···················· ···················· ···················· ···················· ···················· ····················
O 122 hCoV-19/Scotland/QEUH-A5E4A3/2020|EPI_ISL_609182|2  ···················· ···················· ···················· ···················· ···················· ····················
O 123 hCoV-19/Scotland/QEUH-A5DDA9/2020|EPI_ISL_609184|2  ···················· ···················· ···················· ···················· ···················· ····················
O 124 hCoV-19/Wales/QEUH-A5DD9A/2020|EPI_ISL_609185|2020  ···················· ···················· ···················· ···················· ···················· ····················
O 125 hCoV-19/Scotland/QEUH-A5E19D/2020|EPI_ISL_609186|2  ···················· ·A·················· ···················· ···················· ···················· ···A················
O 126 hCoV-19/Scotland/CAMC-A57B69/2020|EPI_ISL_609188|2  ···················· ···················· ···················· ···················· ···················· ····················
O 127 hCoV-19/Wales/QEUH-A5E77D/2020|EPI_ISL_609190|2020  ···················· ···················· ···················· ···················· ···················· ····················
O 128 hCoV-19/Scotland/QEUH-A5E3B5/2020|EPI_ISL_609191|2  ···················· ···················· ···················· ···················· ···················· ····················
O 129 hCoV-19/England/CAMC-A57738/2020|EPI_ISL_609193|20  ···················· ···················· ···················· ···················· ···················· ····················
O 130 hCoV-19/Wales/QEUH-A5DD6D/2020|EPI_ISL_609194|2020  ···················· ···················· ···················· ···················· ···················· ····················
O 131 hCoV-19/Scotland/CAMC-A5764A/2020|EPI_ISL_609195|2  ···················· ·A·······A·········· ···················· ···················· ···················· ····················
O 132 hCoV-19/Scotland/CAMC-A5763B/2020|EPI_ISL_609196|2  ···················· ·A·······A·········· ···················· ···················· ···················· ····················
O 133 hCoV-19/England/QEUH-A5E1BB/2020|EPI_ISL_609197|20  ···················· ···················· ···················· ···················· ···················· ····················
O 134 hCoV-19/Wales/QEUH-A5E4C1/2020|EPI_ISL_609198|2020  ···················· ···················· ···················· ···················· ···················· ····················
O 135 hCoV-19/Scotland/QEUH-A5DF3A/2020|EPI_ISL_609199|2  ···················· ···················· ···················· ···················· ···················· ····················
O 136 hCoV-19/USA/VA-DCLS-1554/2020|EPI_ISL_610118|2020-  ···················· ···················· ···················· ···················· ···················· ····················
O 137 hCoV-19/USA/VA-DCLS-1555/2020|EPI_ISL_610119|2020-  ···················· ···················· ···················· ···················· ···················· ····················
O 138 hCoV-19/USA/VA-DCLS-1556/2020|EPI_ISL_610120|2020-  ···················· ···················· ···················· ···················· ···················· ····················
O 139 hCoV-19/USA/VA-DCLS-1557/2020|EPI_ISL_610121|2020-  ···················· ···················· ···················· ···················· ···················· ····················
O 140 hCoV-19/USA/VA-DCLS-1558/2020|EPI_ISL_610122|2020-  ···················· ······A············· ···················· ···················· ···················· ····················
O 141 hCoV-19/USA/VA-DCLS-1559/2020|EPI_ISL_610123|2020-  ···················· ······A············· ···················· ···················· ···················· ····················
O 142 hCoV-19/USA/VA-DCLS-1560/2020|EPI_ISL_610124|2020-  ···················· ·A·················· ···················· ···················· ···················· ····················
O 143 hCoV-19/USA/VA-DCLS-1561/2020|EPI_ISL_610125|2020-  ···················· ···················· ···················· ···················· ···················· ····················
O 144 hCoV-19/USA/VA-DCLS-1562/2020|EPI_ISL_610126|2020-  ···················· ·A·················· ···················· ···················· ···················· ····················
O 145 hCoV-19/USA/VA-DCLS-1563/2020|EPI_ISL_610127|2020-  ···················· ···················· ···················· ···················· ···················· ····················
O 146 hCoV-19/USA/VA-DCLS-1565/2020|EPI_ISL_610128|2020-  ···················· ·A·················· ···················· ···················· ···················· ····················
O 147 hCoV-19/USA/VA-DCLS-1566/2020|EPI_ISL_610129|2020-  ···················· ···················· ···················· ···················· ···················· ····················
O 148 hCoV-19/USA/VA-DCLS-1567/2020|EPI_ISL_610130|2020-  ···················· ······A············· ···················· ···················· ···················· ····················
O 149 hCoV-19/USA/VA-DCLS-1568/2020|EPI_ISL_610131|2020-  ···················· ···················· ···················· ···················· ···················· ····················
O 150 hCoV-19/USA/VA-DCLS-1571/2020|EPI_ISL_610132|2020-  ···················· ···················· ···················· ···················· ···················· ····················
O 151 hCoV-19/USA/VA-DCLS-1572/2020|EPI_ISL_610133|2020-  ···················· ···················· ···················· ···················· ···················· ····················
O 152 hCoV-19/USA/VA-DCLS-1574/2020|EPI_ISL_610134|2020-  ···················· ···················· ···················· ···················· ···················· ····················
O 153 hCoV-19/USA/VA-DCLS-1575/2020|EPI_ISL_610135|2020-  ···················· ···················· ···················· ···················· ···················· ····················
O 154 hCoV-19/USA/VA-DCLS-1576/2020|EPI_ISL_610136|2020-  ···················· ···················· ···················· ···················· ···················· ····················
O 155 hCoV-19/USA/VA-DCLS-1577/2020|EPI_ISL_610137|2020-  ···················· ···················· ···················· ···················· ···················· ····················
O 156 hCoV-19/USA/VA-DCLS-1578/2020|EPI_ISL_610138|2020-  ········T··········· ·A·················· ···················· ···················· ···················· ····················
O 157 hCoV-19/USA/VA-DCLS-1584/2020|EPI_ISL_610139|2020-  ···················· ···················· ···················· ···················· ···················· ····················
O 158 hCoV-19/USA/VA-DCLS-1585/2020|EPI_ISL_610140|2020-  ···················· ·A·················· ···················· ···················· ···················· ····················
O 159 hCoV-19/USA/VA-DCLS-1586/2020|EPI_ISL_610141|2020-  ···················· ···················· ···················· ···················· ···················· ····················
O 160 hCoV-19/USA/VA-DCLS-1587/2020|EPI_ISL_610142|2020-  ···················· ···················· ···················· ···················· ···················· ····················
O 161 hCoV-19/USA/VA-DCLS-1588/2020|EPI_ISL_610143|2020-  ···················· ···················· ···················· ···················· ···················· ····················
O 162 hCoV-19/USA/VA-DCLS-1590/2020|EPI_ISL_610144|2020-  ···················· ····A··············· ···················· ···················· ···················· ····················
O 163 hCoV-19/USA/VA-DCLS-1591/2020|EPI_ISL_610145|2020-  ···················· ···················· ···················· ···················· ···················· ····················
O 164 hCoV-19/USA/VA-DCLS-1592/2020|EPI_ISL_610146|2020-  ···················· ···················· ···················· ···················· ···················· ····················
O 165 hCoV-19/USA/VA-DCLS-1595/2020|EPI_ISL_610148|2020-  ···················· ···················· ···················· ···················· ···················· ····················
O 166 hCoV-19/USA/VA-DCLS-1598/2020|EPI_ISL_610149|2020-  ···················· ···················· ···················· ···················· ···················· ····················
```

| | | | | | | | |
|---|---|---|---|---|---|---|---|
| O | 167 hCoV-19/USA/MO-WUSTL_E/2020\|EPI_ISL_610160\|2020-08 | ···················· | ···················· | ····················· | ···················· | ···················· | ···················· |
| O | 168 hCoV-19/Indonesia/JT-UGM-48660/2020\|EPI_ISL_610161 | ···················· | ····A··············· | ····················· | ···················· | ···················· | ···················· |
| O | 169 hCoV-19/Indonesia/JT-UGM-47964/2020\|EPI_ISL_610162 | ···················· | ···················· | ····················· | ···················· | ···················· | ···················· |
| O | 170 hCoV-19/England/QEUH-A9EDE8/2020\|EPI_ISL_611400\|20 | ···················· | ···················· | ····················· | ···················· | ···················· | ···················· |
| O | 171 hCoV-19/England/QEUH-AA026C/2020\|EPI_ISL_611401\|20 | ···················· | ···················· | ····················· | ···················· | ···················· | ···················· |
| O | 172 hCoV-19/England/QEUH-A9F0EE/2020\|EPI_ISL_611402\|20 | ···················· | ···················· | ····················· | ···················· | ···················· | ···················· |
| O | 173 hCoV-19/England/QEUH-AA027B/2020\|EPI_ISL_611403\|20 | ···················· | ···················· | ····················· | ···················· | ···················· | ···················· |
| O | 174 hCoV-19/England/QEUH-A9F53A/2020\|EPI_ISL_611448\|20 | ···················· | ···················· | ····················· | ···················· | ···················· | ···················· |
| O | 175 hCoV-19/England/QEUH-A9ECBE/2020\|EPI_ISL_611450\|20 | ···················· | ···················· | ····················· | ···················· | ···················· | ···················· |
| O | 176 hCoV-19/England/QEUH-A9B668/2020\|EPI_ISL_611451\|20 | ···················· | ···················· | ····················· | ···················· | ···················· | ···················· |
| O | 177 hCoV-19/England/QEUH-A9FE11/2020\|EPI_ISL_611452\|20 | ···················· | ···················· | ····················· | ···················· | ···················· | ···················· |
| O | 178 hCoV-19/England/QEUH-A9B6D1/2020\|EPI_ISL_611453\|20 | ···················· | ···················· | ····················· | ···················· | ···················· | ···················· |
| O | 179 hCoV-19/England/QEUH-A9FB38/2020\|EPI_ISL_611455\|20 | ···················· | ···················· | ····················· | ···················· | ···················· | ···················· |
| O | 180 hCoV-19/England/QEUH-AA01D8/2020\|EPI_ISL_611457\|20 | ···················· | ···················· | ····················· | ···················· | ···················· | ···················· |
| O | 181 hCoV-19/England/QEUH-A9F497/2020\|EPI_ISL_611458\|20 | ···················· | ···················· | ····················· | ···················· | ···················· | ···················· |
| O | 182 hCoV-19/England/QEUH-AA0220/2020\|EPI_ISL_611460\|20 | ···················· | ···················· | ····················· | ···················· | ···················· | ···················· |
| O | 183 hCoV-19/England/QEUH-A9E8D8/2020\|EPI_ISL_611461\|20 | ···················· | ···················· | ····················· | ···················· | ···················· | ···················· |
| O | 184 hCoV-19/England/QEUH-AA01C9/2020\|EPI_ISL_611462\|20 | ···················· | ···················· | ····················· | ···················· | ···················· | ···················· |
| O | 185 hCoV-19/England/QEUH-A9F44C/2020\|EPI_ISL_611464\|20 | ···················· | ···················· | ····················· | ···················· | ···················· | ···················· |
| O | 186 hCoV-19/England/QEUH-A9FE7B/2020\|EPI_ISL_611465\|20 | ···················· | ···················· | ····················· | ···················· | ···················· | ···················· |
| O | 187 hCoV-19/England/QEUH-A9E93F/2020\|EPI_ISL_611466\|20 | ···················· | ···················· | ····················· | ···················· | ···················· | ···················· |
| O | 188 hCoV-19/England/QEUH-A9EB84/2020\|EPI_ISL_611469\|20 | ···················· | ···················· | ····················· | ···················· | ····T··············· | ···················· |
| O | 189 hCoV-19/England/QEUH-A9EF5B/2020\|EPI_ISL_611470\|20 | ···················· | ···················· | ····················· | ···················· | ···················· | ···················· |
| O | 190 hCoV-19/England/QEUH-AA019C/2020\|EPI_ISL_611471\|20 | ···················· | ···················· | ····················· | ···················· | ···················· | ···················· |
| O | 191 hCoV-19/England/QEUH-A9E8AB/2020\|EPI_ISL_611472\|20 | ···················· | ···················· | ····················· | ···················· | ···················· | ···················· |
| O | 192 hCoV-19/England/QEUH-AA02A8/2020\|EPI_ISL_611473\|20 | ···················· | ···················· | ····················· | ···················· | ···················· | ···················· |
| O | 193 hCoV-19/England/QEUH-A9ED51/2020\|EPI_ISL_611474\|20 | ···················· | ···················· | ····················· | ···················· | ···················· | ···················· |
| O | 194 hCoV-19/England/QEUH-A9FDF6/2020\|EPI_ISL_611475\|20 | ···················· | ···················· | ····················· | ···················· | ···················· | ···················· |
| O | 195 hCoV-19/England/QEUH-A9ECAF/2020\|EPI_ISL_611477\|20 | ···················· | ···················· | ····················· | ···················· | ···················· | ···················· |
| O | 196 hCoV-19/England/QEUH-A9F655/2020\|EPI_ISL_611478\|20 | ···················· | ···················· | ····················· | ···················· | ···················· | ···················· |
| O | 197 hCoV-19/England/QEUH-A9EE6D/2020\|EPI_ISL_611479\|20 | ···················· | ·A·················· | ····················· | ···················· | ···················· | ···················· |
| O | 198 hCoV-19/USA/UT-UPHL-201029412/2020\|EPI_ISL_611493\| | ···················· | ·A·················· | ····················· | ···················· | ···················· | ···················· |
| O | 199 hCoV-19/USA/UT-UPHL-201003601/2020\|EPI_ISL_611494\| | ···················· | ···················· | ····················· | ···················· | ···················· | ···················· |
| O | 200 hCoV-19/USA/UT-UPHL-201031937/2020\|EPI_ISL_611497\| | ···················· | ···················· | ····················· | ···················· | ···················· | ···················· |
| O | 201 hCoV-19/Wales/PHWC-47F5FA/2020\|EPI_ISL_612700\|2020 | ···················· | ···················· | ····················· | ···················· | ···················· | ···················· |
| O | 202 hCoV-19/Wales/PHWC-47F606/2020\|EPI_ISL_612701\|2020 | ···················· | ···················· | ····················· | ···················· | ···················· | ···················· |
| O | 203 hCoV-19/Wales/PHWC-47F615/2020\|EPI_ISL_612702\|2020 | ···················· | ···················· | ····················· | ···················· | ···················· | ···················· |
| O | 204 hCoV-19/Wales/PHWC-47F624/2020\|EPI_ISL_612703\|2020 | ···················· | ···················· | ····················· | ···················· | ···················· | ···················· |
| O | 205 hCoV-19/Wales/PHWC-47F633/2020\|EPI_ISL_612704\|2020 | ···················· | ···················· | ····················· | ···················· | ···················· | ···················· |
| O | 206 hCoV-19/Wales/PHWC-47F642/2020\|EPI_ISL_612705\|2020 | ···················· | ···················· | ····················· | ···················· | ···················· | ···················· |
| O | 207 hCoV-19/Wales/PHWC-47F651/2020\|EPI_ISL_612706\|2020 | ···················· | ···················· | ····················· | ···················· | ···················· | ···················· |
| O | 208 hCoV-19/Wales/PHWC-47F660/2020\|EPI_ISL_612707\|2020 | ···················· | ···················· | ····················· | ···················· | ···················· | ···················· |
| O | 209 hCoV-19/Wales/PHWC-47F67F/2020\|EPI_ISL_612708\|2020 | ···················· | ···················· | ····················· | ···················· | ···················· | ···················· |
| O | 210 hCoV-19/Wales/PHWC-47F68E/2020\|EPI_ISL_612709\|2020 | ···················· | ···················· | ····················· | ···················· | ···················· | ···················· |
| O | 211 hCoV-19/Wales/PHWC-47F69D/2020\|EPI_ISL_612710\|2020 | ···················· | ···················· | ····················· | ···················· | ···················· | ···················· |
| O | 212 hCoV-19/Wales/PHWC-47F6AC/2020\|EPI_ISL_612711\|2020 | ···················· | ···················· | ····················· | ···················· | ···················· | ···················· |
| O | 213 hCoV-19/Wales/PHWC-47F6CA/2020\|EPI_ISL_612712\|2020 | ···················· | ···················· | ····················· | ···················· | ···················· | ···················· |
| O | 214 hCoV-19/Wales/PHWC-47F6D9/2020\|EPI_ISL_612713\|2020 | ···················· | ···················· | ····················· | ···················· | ···················· | ···················· |
| O | 215 hCoV-19/Wales/PHWC-47F6E8/2020\|EPI_ISL_612714\|2020 | ···················· | ···················· | ····················· | ···················· | ···················· | ···················· |
| O | 216 hCoV-19/Wales/PHWC-47F6F7/2020\|EPI_ISL_612715\|2020 | ···················· | ···················· | ····················· | ···················· | ···················· | ···················· |
| O | 218 hCoV-19/Wales/PHWC-47F721/2020\|EPI_ISL_612717\|2020 | ···················· | ···················· | ····················· | ···················· | ···················· | ···················· |
| O | 219 hCoV-19/Wales/PHWC-47F730/2020\|EPI_ISL_612718\|2020 | ···················· | ···················· | ····················· | ···················· | ···················· | ···················· |
| O | 220 hCoV-19/Wales/PHWC-47F74F/2020\|EPI_ISL_612719\|2020 | ···················· | ···················· | ····················· | ···················· | ···················· | ···················· |
| O | 221 hCoV-19/Wales/PHWC-47F75E/2020\|EPI_ISL_612720\|2020 | ···················· | ···················· | ····················· | ···················· | ···················· | ···················· |
| O | 222 hCoV-19/Wales/PHWC-47F76D/2020\|EPI_ISL_612721\|2020 | ···················· | ···················· | ····················· | ···················· | ···················· | ···················· |
| O | 223 hCoV-19/Wales/PHWC-47F77C/2020\|EPI_ISL_612722\|2020 | ···················· | ···················· | ····················· | ···················· | ···················· | ···················· |
| O | 225 hCoV-19/Wales/PHWC-47F7A9/2020\|EPI_ISL_612724\|2020 | ···················· | ···················· | ····················· | ···················· | ···················· | ···················· |
| O | 226 hCoV-19/Wales/PHWC-47F7C7/2020\|EPI_ISL_612725\|2020 | ···················· | ···················· | ····················· | ···················· | ···················· | ···················· |
| O | 227 hCoV-19/Wales/PHWC-47F7D6/2020\|EPI_ISL_612726\|2020 | ···················· | ···················· | ····················· | ···················· | ···················· | ···················· |
| O | 228 hCoV-19/Wales/PHWC-47F7E5/2020\|EPI_ISL_612727\|2020 | ···················· | ···················· | ····················· | ···················· | ···················· | ···················· |
| O | 229 hCoV-19/Wales/PHWC-47F7F4/2020\|EPI_ISL_612728\|2020 | ···················· | ···················· | ····················· | ···················· | ···················· | ···················· |
| O | 230 hCoV-19/Wales/PHWC-47F800/2020\|EPI_ISL_612729\|2020 | ···················· | ···················· | ····················· | ···················· | ···················· | ···················· |
| O | 231 hCoV-19/Wales/PHWC-47F81F/2020\|EPI_ISL_612730\|2020 | ···················· | ···················· | ····················· | ···················· | ···················· | ···················· |
| O | 232 hCoV-19/Wales/PHWC-47F82E/2020\|EPI_ISL_612731\|2020 | ···················· | ···················· | ····················· | ···················· | ···················· | ···················· |
| O | 233 hCoV-19/Wales/PHWC-47F83D/2020\|EPI_ISL_612732\|2020 | ···················· | ···················· | ····················· | ···················· | ···················· | ···················· |
| O | 234 hCoV-19/Wales/PHWC-47F84C/2020\|EPI_ISL_612733\|2020 | ···················· | ···················· | ····················· | ···················· | ···················· | ···················· |
| O | 235 hCoV-19/Wales/PHWC-47F85B/2020\|EPI_ISL_612734\|2020 | ···················· | ···················· | ····················· | ···················· | ···················· | ···················· |
| O | 236 hCoV-19/Wales/PHWC-47FB8F/2020\|EPI_ISL_612779\|2020 | ···················· | ···················· | ····················· | ···················· | ···················· | ···················· |
| O | 237 hCoV-19/Wales/PHWC-47FB9E/2020\|EPI_ISL_612780\|2020 | ···················· | ···················· | ····················· | ···················· | ···················· | ···················· |
| O | 238 hCoV-19/Wales/PHWC-47FBAD/2020\|EPI_ISL_612781\|2020 | ···················· | ···················· | ····················· | ···················· | ···················· | ···················· |
| O | 239 hCoV-19/Wales/PHWC-47FBBC/2020\|EPI_ISL_612782\|2020 | ···················· | ···················· | ····················· | ···················· | ···················· | ···················· |
| O | 240 hCoV-19/Wales/PHWC-47FBCB/2020\|EPI_ISL_612783\|2020 | ···················· | ···················· | ····················· | ···················· | ···················· | ···················· |
| O | 241 hCoV-19/Wales/PHWC-47FBDA/2020\|EPI_ISL_612784\|2020 | ···················· | ···················· | ····················· | ···················· | ···················· | ···················· |
| O | 242 hCoV-19/Wales/PHWC-47FBF8/2020\|EPI_ISL_612785\|2020 | ···················· | ···················· | ····················· | ···················· | ···················· | ···················· |
| O | 243 hCoV-19/Wales/PHWC-47FC04/2020\|EPI_ISL_612786\|2020 | ···················· | ···················· | ····················· | ···················· | ···················· | ···················· |
| O | 244 hCoV-19/Wales/PHWC-47FC13/2020\|EPI_ISL_612787\|2020 | ···················· | ···················· | ····················· | ···················· | ···················· | ···················· |
| O | 245 hCoV-19/Wales/PHWC-47FC22/2020\|EPI_ISL_612788\|2020 | ···················· | ···················· | ····················· | ···················· | ···················· | ···················· |
| O | 248 hCoV-19/Wales/PHWC-47FC5F/2020\|EPI_ISL_612791\|2020 | ···················· | ···················· | ····················· | ···················· | ···················· | ···················· |
| O | 249 hCoV-19/Wales/PHWC-47FC6E/2020\|EPI_ISL_612792\|2020 | ···················· | ···················· | ····················· | ···················· | ···················· | ···················· |
| O | 250 hCoV-19/Wales/PHWC-47FC7D/2020\|EPI_ISL_612793\|2020 | ···················· | ···················· | ····················· | ···················· | ···················· | ···················· |

| | | | | | | | |
|---|---|---|---|---|---|---|---|
| O | 251 hCoV-19/Wales/PHWC-47FC8C/2020\|EPI_ISL_612794\|2020 | .................... | .................... | .................... | .................... | .................... | .................... |
| O | 252 hCoV-19/Wales/PHWC-47FC9B/2020\|EPI_ISL_612795\|2020 | .................... | .................... | .................... | .................... | .................... | .................... |
| O | 253 hCoV-19/Wales/PHWC-47FCAA/2020\|EPI_ISL_612796\|2020 | .................... | .................... | .................... | .................... | .................... | .................... |
| O | 254 hCoV-19/Wales/PHWC-47FCB9/2020\|EPI_ISL_612797\|2020 | .................... | .................... | .................... | .................... | .................... | .................... |
| O | 255 hCoV-19/Wales/PHWC-47FCC8/2020\|EPI_ISL_612798\|2020 | .................... | .................... | .................... | .................... | .................... | .................... |
| O | 256 hCoV-19/Wales/PHWC-47FCD7/2020\|EPI_ISL_612799\|2020 | .................... | .................... | .................... | .................... | .................... | .................... |
| O | 257 hCoV-19/England/MILK-A7A113/2020\|EPI_ISL_606817\|20 | .................... | .................... | .................... | .................... | .................... | .................... |
| O | 258 hCoV-19/England/MILK-A79949/2020\|EPI_ISL_606818\|20 | .................... | .A.................. | .................... | .................... | .................... | .................... |
| O | 259 hCoV-19/England/MILK-A7AC5E/2020\|EPI_ISL_606819\|20 | .................... | .................... | .................... | .................... | .................... | .................... |
| O | 260 hCoV-19/England/MILK-A7A5AE/2020\|EPI_ISL_606820\|20 | .................... | .................... | .................... | .................... | .................... | .................... |
| O | 261 hCoV-19/England/MILK-A7D08C/2020\|EPI_ISL_606822\|20 | .................... | .................... | .................... | .................... | .................... | .................... |
| O | 262 hCoV-19/England/MILK-A79E0E/2020\|EPI_ISL_606823\|20 | .................... | .................... | .................... | .................... | .................... | .................... |
| O | 263 hCoV-19/England/MILK-A7A614/2020\|EPI_ISL_606824\|20 | .................... | .................... | .................... | .................... | .................... | .................... |
| O | 264 hCoV-19/England/MILK-A79A19/2020\|EPI_ISL_606825\|20 | .................... | .................... | .................... | .................... | .................... | .................... |
| O | 265 hCoV-19/England/MILK-A7A5EA/2020\|EPI_ISL_606826\|20 | .................... | .................... | .................... | .................... | .................... | .................... |
| O | 266 hCoV-19/England/MILK-A7CF44/2020\|EPI_ISL_606827\|20 | .................... | .A.................. | .................... | .................... | .................... | .................... |
| O | 267 hCoV-19/England/MILK-A79E1D/2020\|EPI_ISL_606828\|20 | .................... | .................... | .................... | .................... | .................... | .................... |
| O | 268 hCoV-19/England/MILK-A7AD4C/2020\|EPI_ISL_606830\|20 | .................... | .................... | .................... | .................... | .................... | .................... |
| O | 269 hCoV-19/England/MILK-A7A720/2020\|EPI_ISL_606831\|20 | .................... | .................... | .................... | .................... | .................... | .................... |
| O | 270 hCoV-19/England/MILK-A79D89/2020\|EPI_ISL_606832\|20 | .................... | .................... | .................... | .................... | .................... | .................... |
| O | 271 hCoV-19/England/MILK-A79CF5/2020\|EPI_ISL_606834\|20 | .................... | .................... | .................... | .................... | .................... | .................... |
| O | 272 hCoV-19/England/MILK-A7CF35/2020\|EPI_ISL_606835\|20 | .................... | .................... | .................... | .................... | .................... | .................... |
| O | 273 hCoV-19/England/MILK-A79A82/2020\|EPI_ISL_606836\|20 | .................... | .................... | .................... | .................... | .................... | .................... |
| O | 274 hCoV-19/England/MILK-A79B61/2020\|EPI_ISL_606837\|20 | .................... | .................... | .................... | .................... | .................... | .................... |
| O | 275 hCoV-19/England/MILK-A7A2B6/2020\|EPI_ISL_606838\|20 | .................... | .................... | .................... | .................... | .................... | .................... |
| O | 276 hCoV-19/England/MILK-A79976/2020\|EPI_ISL_606839\|20 | .................... | .................... | .................... | .................... | .................... | .................... |
| O | 277 hCoV-19/England/MILK-A79590/2020\|EPI_ISL_606840\|20 | .................... | .................... | .................... | .................... | .................... | .................... |
| O | 278 hCoV-19/England/MILK-A7ACC7/2020\|EPI_ISL_606841\|20 | .................... | .................... | .................... | .................... | .................... | .................... |
| O | 279 hCoV-19/England/MILK-A79572/2020\|EPI_ISL_606842\|20 | .................... | .................... | .................... | .................... | .................... | .................... |
| O | 280 hCoV-19/England/MILK-A7A474/2020\|EPI_ISL_606843\|20 | .................... | .................... | .................... | .................... | .................... | .................... |
| O | 281 hCoV-19/England/MILK-A796BB/2020\|EPI_ISL_606846\|20 | .................... | .................... | .................... | .................... | .................... | .................... |
| O | 282 hCoV-19/England/MILK-A7A052/2020\|EPI_ISL_606847\|20 | .................... | .................... | .................... | .................... | .................... | .................... |
| O | 283 hCoV-19/England/MILK-A79721/2020\|EPI_ISL_606848\|20 | .................... | .................... | .................... | .................... | ......T............. | .................... |
| O | 284 hCoV-19/England/MILK-A7CEB0/2020\|EPI_ISL_606849\|20 | .................... | .................... | .................... | .................... | .................... | .................... |
| O | 285 hCoV-19/England/MILK-A7A386/2020\|EPI_ISL_606850\|20 | .................... | .................... | .................... | .................... | ......T............. | .................... |
| O | 286 hCoV-19/England/MILK-A7A08F/2020\|EPI_ISL_606851\|20 | .................... | .................... | .................... | .................... | .................... | .................... |
| O | 287 hCoV-19/England/MILK-A798F1/2020\|EPI_ISL_606852\|20 | .................... | .................... | .................... | .................... | .................... | .................... |
| O | 288 hCoV-19/England/MILK-A7A0E9/2020\|EPI_ISL_606853\|20 | .................... | .................... | .................... | .................... | .................... | .................... |
| O | 289 hCoV-19/England/MILK-A79994/2020\|EPI_ISL_606854\|20 | .................... | .................... | .................... | .................... | .................... | .................... |
| O | 290 hCoV-19/England/MILK-A7A18C/2020\|EPI_ISL_606855\|20 | .................... | .................... | .................... | .................... | .................... | .................... |
| O | 291 hCoV-19/England/MILK-A7CDE0/2020\|EPI_ISL_606856\|20 | .................... | .................... | .................... | .................... | .................... | .................... |
| O | 292 hCoV-19/England/MILK-A7AB7F/2020\|EPI_ISL_606857\|20 | .................... | .................... | .................... | .................... | ......T............. | .................... |
| O | 293 hCoV-19/England/MILK-A79E2C/2020\|EPI_ISL_606858\|20 | .................... | .................... | .................... | .................... | .................... | .................... |
| O | 294 hCoV-19/England/MILK-A79B34/2020\|EPI_ISL_606859\|20 | .................... | .................... | .................... | .................... | .................... | .................... |
| O | 295 hCoV-19/England/CAMC-A6504B/2020\|EPI_ISL_609050\|20 | .................... | .................... | .................... | .................... | .................... | .................... |
| O | 296 hCoV-19/England/ALDP-A695B1/2020\|EPI_ISL_609051\|20 | .................... | .................... | .................... | .................... | .................... | .................... |
| O | 297 hCoV-19/England/ALDP-A68FE1/2020\|EPI_ISL_609052\|20 | .................... | .................... | .................... | .................... | .................... | .................... |
| O | 298 hCoV-19/England/ALDP-A680B1/2020\|EPI_ISL_609053\|20 | .................... | .................... | .................... | .................... | .................... | .................... |
| O | 299 hCoV-19/England/CAMC-A64A7B/2020\|EPI_ISL_609054\|20 | .................... | .................... | .................... | .................... | .................... | .................... |
| O | 300 hCoV-19/England/ALDP-A67F97/2020\|EPI_ISL_609055\|20 | .................... | .................... | .................... | .................... | .................... | .................... |
| O | 301 hCoV-19/England/ALDP-A68A1D/2020\|EPI_ISL_609056\|20 | .................... | .................... | .................... | .................... | .................... | .................... |
| O | 302 hCoV-19/England/ALDP-A682AC/2020\|EPI_ISL_609057\|20 | .................... | .................... | .................... | .................... | .................... | ...A................ |
| O | 303 hCoV-19/England/MILK-A6BFFD/2020\|EPI_ISL_609058\|20 | .................... | .................... | .................... | .................... | .................... | .................... |
| O | 304 hCoV-19/England/MILK-A6BE4B/2020\|EPI_ISL_609059\|20 | .................... | .................... | .................... | .................... | .................... | .................... |
| O | 305 hCoV-19/England/ALDP-A680C0/2020\|EPI_ISL_609060\|20 | .................... | .................... | .................... | .................... | .................... | .................... |
| O | 306 hCoV-19/England/MILK-A6BEF0/2020\|EPI_ISL_609062\|20 | .................... | .................... | .................... | .................... | .................... | .................... |
| O | 307 hCoV-19/England/MILK-A6BDB7/2020\|EPI_ISL_609063\|20 | .................... | .................... | .................... | .................... | .................... | .................... |
| O | 308 hCoV-19/England/ALDP-A695EE/2020\|EPI_ISL_609064\|20 | .................... | .................... | .................... | .................... | .................... | .................... |
| O | 309 hCoV-19/England/ALDP-A69584/2020\|EPI_ISL_609065\|20 | .................... | .................... | .................... | .................... | .................... | .................... |
| O | 310 hCoV-19/England/MILK-A6C1AB/2020\|EPI_ISL_609066\|20 | .................... | .................... | .................... | .................... | .................... | .................... |
| O | 311 hCoV-19/England/ALDP-A6940F/2020\|EPI_ISL_609068\|20 | .................... | .................... | .................... | .................... | .................... | .................... |
| O | 312 hCoV-19/England/ALDP-A695C0/2020\|EPI_ISL_609075\|20 | .................... | .................... | .................... | .................... | .................... | .................... |
| O | 313 hCoV-19/England/MILK-A6BFA2/2020\|EPI_ISL_609076\|20 | .................... | .................... | .................... | .................... | .................... | .................... |
| O | 314 hCoV-19/England/CAMC-A64905/2020\|EPI_ISL_609077\|20 | .................... | .................... | .................... | .................... | .................... | .................... |
| O | 315 hCoV-19/England/CAMC-A646D1/2020\|EPI_ISL_609091\|20 | .................... | .................... | .................... | .................... | .................... | .................... |
| O | 316 hCoV-19/England/ALDP-A69548/2020\|EPI_ISL_609093\|20 | .................... | .................... | .................... | .................... | .................... | .................... |
| O | 317 hCoV-19/England/ALDP-A69205/2020\|EPI_ISL_609095\|20 | .................... | .................... | .................... | .................... | .................... | .................... |
| O | 318 hCoV-19/England/CAMC-A64747/2020\|EPI_ISL_609097\|20 | .................... | .................... | .................... | .................... | .................... | .................... |
| O | 319 hCoV-19/England/ALDP-A6834F/2020\|EPI_ISL_609098\|20 | .................... | .................... | .................... | .................... | .................... | .................... |
| O | 320 hCoV-19/England/CAMC-A646FF/2020\|EPI_ISL_609099\|20 | .................... | .................... | .................... | .................... | .................... | .................... |
| O | 321 hCoV-19/England/CAMC-A646C2/2020\|EPI_ISL_609100\|20 | .................... | .................... | .................... | .................... | .................... | .................... |
| O | 322 hCoV-19/England/CAMC-A6471A/2020\|EPI_ISL_609101\|20 | .................... | .................... | .................... | .................... | .................... | .................... |
| O | 323 hCoV-19/England/ALDP-A6952A/2020\|EPI_ISL_609102\|20 | .................... | .................... | .................... | .................... | .................... | .................... |
| O | 324 hCoV-19/England/ALDP-A68127/2020\|EPI_ISL_609103\|20 | .................... | .................... | .................... | .................... | .................... | .................... |
| O | 325 hCoV-19/England/ALDP-A68B1A/2020\|EPI_ISL_609104\|20 | .................... | .................... | .................... | .................... | .................... | .................... |
| O | 326 hCoV-19/England/ALDP-A6927E/2020\|EPI_ISL_609105\|20 | .................... | .................... | .................... | .................... | .................... | .................... |
| O | 327 hCoV-19/England/ALDP-A67DAC/2020\|EPI_ISL_609107\|20 | .................... | .................... | .................... | .................... | .................... | .................... |
| O | 328 hCoV-19/England/MILK-A6BE87/2020\|EPI_ISL_609108\|20 | .................... | .................... | .................... | .................... | .................... | .................... |
| O | 329 hCoV-19/England/ALDP-A68260/2020\|EPI_ISL_609109\|20 | .................... | .................... | .................... | .................... | .................... | .................... |
| O | 330 hCoV-19/England/MILK-A6BF39/2020\|EPI_ISL_609110\|20 | .................... | .................... | .................... | .................... | .................... | .................... |

<table>
<tr><td>O</td><td>331</td><td>hCoV-19/England/CAMC-A64835/2020|EPI_ISL_609111|20</td><td>····················</td><td>····················</td><td>····················</td><td>····················</td><td>····················</td><td>····················</td></tr>
<tr><td>O</td><td>332</td><td>hCoV-19/England/CAMC-A645E3/2020|EPI_ISL_609113|20</td><td>····················</td><td>····················</td><td>····················</td><td>····················</td><td>····················</td><td>····················</td></tr>
<tr><td>O</td><td>333</td><td>hCoV-19/England/CAMC-A64941/2020|EPI_ISL_609114|20</td><td>····················</td><td>····················</td><td>····················</td><td>····················</td><td>····················</td><td>····················</td></tr>
<tr><td>O</td><td>334</td><td>hCoV-19/England/MILK-A6C017/2020|EPI_ISL_609115|20</td><td>····················</td><td>·A·······A··········</td><td>····················</td><td>····················</td><td>····················</td><td>····················</td></tr>
<tr><td>O</td><td>335</td><td>hCoV-19/England/MILK-A6BF57/2020|EPI_ISL_609116|20</td><td>·····G··············</td><td>····················</td><td>····················</td><td>····················</td><td>····················</td><td>····················</td></tr>
<tr><td>O</td><td>336</td><td>hCoV-19/England/MILK-A6C18D/2020|EPI_ISL_609117|20</td><td>····················</td><td>····················</td><td>····················</td><td>····················</td><td>····················</td><td>····················</td></tr>
<tr><td>O</td><td>337</td><td>hCoV-19/England/CAMC-A64844/2020|EPI_ISL_609118|20</td><td>····················</td><td>····················</td><td>····················</td><td>····················</td><td>····················</td><td>····················</td></tr>
<tr><td>O</td><td>338</td><td>hCoV-19/England/ALDP-A690DE/2020|EPI_ISL_609120|20</td><td>····················</td><td>····················</td><td>····················</td><td>····················</td><td>····················</td><td>····················</td></tr>
<tr><td>O</td><td>339</td><td>hCoV-19/England/CAMC-A64AA8/2020|EPI_ISL_609122|20</td><td>····················</td><td>····················</td><td>····················</td><td>····················</td><td>····················</td><td>····················</td></tr>
<tr><td>O</td><td>340</td><td>hCoV-19/England/CAMC-A64659/2020|EPI_ISL_609123|20</td><td>····················</td><td>····················</td><td>····················</td><td>····················</td><td>····················</td><td>····················</td></tr>
<tr><td>O</td><td>341</td><td>hCoV-19/England/ALDP-A6938A/2020|EPI_ISL_609124|20</td><td>····················</td><td>····················</td><td>····················</td><td>····················</td><td>····················</td><td>····················</td></tr>
<tr><td>O</td><td>342</td><td>hCoV-19/England/MILK-A6BFB1/2020|EPI_ISL_609126|20</td><td>····················</td><td>····················</td><td>····················</td><td>····················</td><td>····················</td><td>····················</td></tr>
<tr><td>O</td><td>343</td><td>hCoV-19/England/ALDP-A690A1/2020|EPI_ISL_609127|20</td><td>····················</td><td>····················</td><td>····················</td><td>····················</td><td>····················</td><td>····················</td></tr>
<tr><td>O</td><td>344</td><td>hCoV-19/England/MILK-A6C0AE/2020|EPI_ISL_609128|20</td><td>····················</td><td>····················</td><td>····················</td><td>····················</td><td>····················</td><td>····················</td></tr>
<tr><td>O</td><td>345</td><td>hCoV-19/England/CAMC-A648CC/2020|EPI_ISL_609129|20</td><td>····················</td><td>····················</td><td>····················</td><td>····················</td><td>····················</td><td>····················</td></tr>
<tr><td>O</td><td>346</td><td>hCoV-19/England/CAMC-A59262/2020|EPI_ISL_609160|20</td><td>····················</td><td>····················</td><td>····················</td><td>····················</td><td>····················</td><td>····················</td></tr>
<tr><td>O</td><td>347</td><td>hCoV-19/Wales/CAMC-A58A6B/2020|EPI_ISL_609161|2020</td><td>····················</td><td>····················</td><td>····················</td><td>····················</td><td>····················</td><td>····················</td></tr>
<tr><td>O</td><td>348</td><td>hCoV-19/Wales/CAMC-A58AF2/2020|EPI_ISL_609162|2020</td><td>····················</td><td>····················</td><td>····················</td><td>····················</td><td>····················</td><td>····················</td></tr>
<tr><td>O</td><td>349</td><td>hCoV-19/Wales/CAMC-A590E0/2020|EPI_ISL_609163|2020</td><td>····················</td><td>····················</td><td>····················</td><td>····················</td><td>····················</td><td>····················</td></tr>
<tr><td>O</td><td>350</td><td>hCoV-19/Wales/CAMC-A595A5/2020|EPI_ISL_609164|2020</td><td>····················</td><td>····················</td><td>····················</td><td>····················</td><td>····················</td><td>····················</td></tr>
<tr><td>O</td><td>351</td><td>hCoV-19/Wales/CAMC-A5811B/2020|EPI_ISL_609165|2020</td><td>····················</td><td>····················</td><td>····················</td><td>····················</td><td>····················</td><td>····················</td></tr>
<tr><td>O</td><td>352</td><td>hCoV-19/Wales/CAMC-A58E9C/2020|EPI_ISL_609166|2020</td><td>····················</td><td>····················</td><td>····················</td><td>····················</td><td>····················</td><td>····················</td></tr>
<tr><td>O</td><td>353</td><td>hCoV-19/Scotland/CAMC-A588BC/2020|EPI_ISL_609167|2</td><td>····················</td><td>····················</td><td>····················</td><td>····················</td><td>····················</td><td>····················</td></tr>
<tr><td>O</td><td>354</td><td>hCoV-19/Scotland/CAMC-A58728/2020|EPI_ISL_609168|2</td><td>····················</td><td>····················</td><td>····················</td><td>····················</td><td>····················</td><td>····················</td></tr>
<tr><td>O</td><td>355</td><td>hCoV-19/Wales/CAMC-A58351/2020|EPI_ISL_609169|2020</td><td>····················</td><td>····················</td><td>····················</td><td>····················</td><td>····················</td><td>····················</td></tr>
<tr><td>O</td><td>356</td><td>hCoV-19/USA/TX-DSHS-0909/2020|EPI_ISL_610045|2020-</td><td>····················</td><td>····················</td><td>····················</td><td>····················</td><td>N···················</td><td>····················</td></tr>
<tr><td>O</td><td>357</td><td>hCoV-19/USA/MI-UM-10036399817/2020|EPI_ISL_610048|</td><td>····················</td><td>····················</td><td>····················</td><td>····················</td><td>····················</td><td>····················</td></tr>
<tr><td>O</td><td>358</td><td>hCoV-19/USA/MI-UM-10036457442/2020|EPI_ISL_610049|</td><td>····················</td><td>····················</td><td>····················</td><td>····················</td><td>····················</td><td>····················</td></tr>
<tr><td>O</td><td>359</td><td>hCoV-19/USA/MI-UM-10036399786/2020|EPI_ISL_610050|</td><td>····················</td><td>····················</td><td>····················</td><td>····················</td><td>····················</td><td>····················</td></tr>
<tr><td>O</td><td>360</td><td>hCoV-19/USA/MI-UM-10036477051/2020|EPI_ISL_610051|</td><td>····················</td><td>····················</td><td>····················</td><td>····················</td><td>····················</td><td>····················</td></tr>
<tr><td>O</td><td>361</td><td>hCoV-19/USA/MI-UM-10036457372/2020|EPI_ISL_610052|</td><td>····················</td><td>····················</td><td>····················</td><td>····················</td><td>····················</td><td>····················</td></tr>
<tr><td>O</td><td>363</td><td>hCoV-19/USA/MI-UM-10036345655/2020|EPI_ISL_610054|</td><td>····················</td><td>····················</td><td>····················</td><td>····················</td><td>····················</td><td>····················</td></tr>
<tr><td>O</td><td>367</td><td>hCoV-19/USA/MI-UM-10036335706/2020|EPI_ISL_610059|</td><td>····················</td><td>····················</td><td>····················</td><td>····················</td><td>····················</td><td>····················</td></tr>
<tr><td>O</td><td>368</td><td>hCoV-19/USA/MI-UM-10036392191/2020|EPI_ISL_610065|</td><td>····················</td><td>····················</td><td>····················</td><td>····················</td><td>····················</td><td>····················</td></tr>
<tr><td>O</td><td>369</td><td>hCoV-19/USA/MI-UM-10036396766/2020|EPI_ISL_610066|</td><td>····················</td><td>····················</td><td>····················</td><td>····················</td><td>····················</td><td>····················</td></tr>
<tr><td>O</td><td>370</td><td>hCoV-19/USA/MI-UM-10036322730/2020|EPI_ISL_610067|</td><td>····················</td><td>····················</td><td>····················</td><td>····················</td><td>····················</td><td>····················</td></tr>
<tr><td>O</td><td>371</td><td>hCoV-19/USA/MI-UM-10036394525/2020|EPI_ISL_610068|</td><td>····················</td><td>····················</td><td>····················</td><td>····················</td><td>····················</td><td>····················</td></tr>
<tr><td>O</td><td>372</td><td>hCoV-19/USA/MI-UM-10036415960/2020|EPI_ISL_610083|</td><td>····················</td><td>····················</td><td>····················</td><td>····················</td><td>····················</td><td>····················</td></tr>
<tr><td>O</td><td>373</td><td>hCoV-19/USA/MI-UM-10036396778/2020|EPI_ISL_610084|</td><td>····················</td><td>····················</td><td>····················</td><td>····················</td><td>····················</td><td>····················</td></tr>
<tr><td>O</td><td>374</td><td>hCoV-19/USA/MI-UM-10036396939/2020|EPI_ISL_610085|</td><td>····················</td><td>····················</td><td>····················</td><td>····················</td><td>····················</td><td>····················</td></tr>
<tr><td>O</td><td>378</td><td>hCoV-19/USA/MI-UM-10036416305/2020|EPI_ISL_610089|</td><td>····················</td><td>····················</td><td>····················</td><td>····················</td><td>····················</td><td>····················</td></tr>
<tr><td>O</td><td>383</td><td>hCoV-19/USA/MI-UM-10036429589/2020|EPI_ISL_610095|</td><td>····················</td><td>····················</td><td>····················</td><td>····················</td><td>····················</td><td>····················</td></tr>
<tr><td>O</td><td>385</td><td>hCoV-19/USA/MI-UM-10036448224/2020|EPI_ISL_610101|</td><td>····················</td><td>····················</td><td>····················</td><td>····················</td><td>····················</td><td>····················</td></tr>
<tr><td>O</td><td>387</td><td>hCoV-19/USA/MI-UM-10036424368/2020|EPI_ISL_610104|</td><td>····················</td><td>·A··················</td><td>····················</td><td>····················</td><td>····················</td><td>····················</td></tr>
<tr><td>O</td><td>393</td><td>hCoV-19/USA/MI-UM-10036438245/2020|EPI_ISL_610111|</td><td>····················</td><td>····················</td><td>····················</td><td>····················</td><td>····················</td><td>····················</td></tr>
<tr><td>O</td><td>396</td><td>hCoV-19/USA/VA-DCLS-1599/2020|EPI_ISL_610150|2020-</td><td>····················</td><td>····················</td><td>····················</td><td>····················</td><td>····················</td><td>····················</td></tr>
<tr><td>O</td><td>397</td><td>hCoV-19/USA/VA-DCLS-1600/2020|EPI_ISL_610151|2020-</td><td>····················</td><td>·A··················</td><td>····················</td><td>····················</td><td>····················</td><td>····················</td></tr>
<tr><td>O</td><td>398</td><td>hCoV-19/Singapore/1110nan/2020|EPI_ISL_610152|2020</td><td>····················</td><td>····················</td><td>····················</td><td>····················</td><td>····················</td><td>····················</td></tr>
<tr><td>O</td><td>399</td><td>hCoV-19/Singapore/1083nan/2020|EPI_ISL_610153|2020</td><td>····················</td><td>····················</td><td>····················</td><td>····················</td><td>····················</td><td>····················</td></tr>
<tr><td>O</td><td>400</td><td>hCoV-19/Singapore/1116/2020|EPI_ISL_610154|2020-10</td><td>····················</td><td>····················</td><td>····················</td><td>····················</td><td>····················</td><td>····················</td></tr>
<tr><td>O</td><td>401</td><td>hCoV-19/USA/MO-WUSTL_C/2020|EPI_ISL_610157|2020-08</td><td>····················</td><td>····················</td><td>····················</td><td>····················</td><td>····················</td><td>····················</td></tr>
<tr><td>O</td><td>402</td><td>hCoV-19/USA/MO-WUSTL_D/2020|EPI_ISL_610159|2020-08</td><td>····················</td><td>····················</td><td>····················</td><td>····················</td><td>····················</td><td>····················</td></tr>
<tr><td>O</td><td>403</td><td>hCoV-19/England/QEUH-A9F118/2020|EPI_ISL_611370|20</td><td>····················</td><td>····················</td><td>····················</td><td>····················</td><td>····················</td><td>····················</td></tr>
<tr><td>O</td><td>404</td><td>hCoV-19/England/QEUH-A9B6C2/2020|EPI_ISL_611371|20</td><td>····················</td><td>····················</td><td>····················</td><td>····················</td><td>····················</td><td>····················</td></tr>
<tr><td>O</td><td>405</td><td>hCoV-19/England/QEUH-AA02F3/2020|EPI_ISL_611372|20</td><td>····················</td><td>····················</td><td>····················</td><td>····················</td><td>····················</td><td>····················</td></tr>
<tr><td>O</td><td>406</td><td>hCoV-19/England/QEUH-A9E8C9/2020|EPI_ISL_611373|20</td><td>····················</td><td>····················</td><td>····················</td><td>····················</td><td>····················</td><td>····················</td></tr>
<tr><td>O</td><td>407</td><td>hCoV-19/England/QEUH-AA03E1/2020|EPI_ISL_611374|20</td><td>····················</td><td>····················</td><td>····················</td><td>····················</td><td>····················</td><td>····················</td></tr>
<tr><td>O</td><td>408</td><td>hCoV-19/England/QEUH-A9F5D0/2020|EPI_ISL_611375|20</td><td>····················</td><td>····················</td><td>····················</td><td>····················</td><td>····················</td><td>····················</td></tr>
<tr><td>O</td><td>409</td><td>hCoV-19/England/QEUH-A9FDAB/2020|EPI_ISL_611377|20</td><td>····················</td><td>····················</td><td>····················</td><td>····················</td><td>····················</td><td>····················</td></tr>
<tr><td>O</td><td>410</td><td>hCoV-19/England/QEUH-AA0211/2020|EPI_ISL_611379|20</td><td>····················</td><td>····················</td><td>····················</td><td>····················</td><td>····················</td><td>····················</td></tr>
<tr><td>O</td><td>411</td><td>hCoV-19/England/QEUH-A9F707/2020|EPI_ISL_611380|20</td><td>····················</td><td>····················</td><td>····················</td><td>····················</td><td>····················</td><td>····················</td></tr>
<tr><td>O</td><td>412</td><td>hCoV-19/England/QEUH-A9B765/2020|EPI_ISL_611381|20</td><td>····················</td><td>····················</td><td>····················</td><td>····················</td><td>····················</td><td>····················</td></tr>
<tr><td>O</td><td>413</td><td>hCoV-19/England/QEUH-A9EC27/2020|EPI_ISL_611382|20</td><td>····················</td><td>····················</td><td>····················</td><td>····················</td><td>····················</td><td>····················</td></tr>
<tr><td>O</td><td>414</td><td>hCoV-19/England/QEUH-A9F479/2020|EPI_ISL_611383|20</td><td>····················</td><td>····················</td><td>····················</td><td>····················</td><td>····················</td><td>····················</td></tr>
<tr><td>O</td><td>415</td><td>hCoV-19/England/QEUH-A9EED6/2020|EPI_ISL_611384|20</td><td>····················</td><td>····················</td><td>····················</td><td>····················</td><td>····················</td><td>····················</td></tr>
<tr><td>O</td><td>416</td><td>hCoV-19/England/QEUH-A9EEC7/2020|EPI_ISL_611385|20</td><td>····················</td><td>····················</td><td>····················</td><td>····················</td><td>····················</td><td>····················</td></tr>
<tr><td>O</td><td>417</td><td>hCoV-19/England/QEUH-A9F6DD/2020|EPI_ISL_611386|20</td><td>····················</td><td>····················</td><td>····················</td><td>····················</td><td>····················</td><td>····················</td></tr>
<tr><td>O</td><td>418</td><td>hCoV-19/England/QEUH-A9F163/2020|EPI_ISL_611387|20</td><td>····················</td><td>····················</td><td>····················</td><td>····················</td><td>····················</td><td>····················</td></tr>
<tr><td>O</td><td>419</td><td>hCoV-19/England/QEUH-A9FDC9/2020|EPI_ISL_611389|20</td><td>····················</td><td>····················</td><td>····················</td><td>····················</td><td>····················</td><td>····················</td></tr>
<tr><td>O</td><td>420</td><td>hCoV-19/England/QEUH-AA03C3/2020|EPI_ISL_611396|20</td><td>····················</td><td>····················</td><td>····················</td><td>····················</td><td>····················</td><td>····················</td></tr>
<tr><td>O</td><td>421</td><td>hCoV-19/England/QEUH-AA025D/2020|EPI_ISL_611397|20</td><td>····················</td><td>····················</td><td>····················</td><td>····················</td><td>····················</td><td>····················</td></tr>
<tr><td>O</td><td>422</td><td>hCoV-19/England/QEUH-A9EC90/2020|EPI_ISL_611398|20</td><td>····················</td><td>····················</td><td>····················</td><td>····················</td><td>····················</td><td>····················</td></tr>
<tr><td>O</td><td>423</td><td>hCoV-19/England/QEUH-A9EE8B/2020|EPI_ISL_611404|20</td><td>····················</td><td>····················</td><td>····················</td><td>····················</td><td>····················</td><td>····················</td></tr>
<tr><td>O</td><td>424</td><td>hCoV-19/England/QEUH-A9E902/2020|EPI_ISL_611405|20</td><td>····················</td><td>····················</td><td>····················</td><td>····················</td><td>····················</td><td>····················</td></tr>
<tr><td>O</td><td>425</td><td>hCoV-19/England/QEUH-A9FBCF/2020|EPI_ISL_611406|20</td><td>····················</td><td>····················</td><td>····················</td><td>····················</td><td>····················</td><td>····················</td></tr>
<tr><td>O</td><td>426</td><td>hCoV-19/England/QEUH-A9B71A/2020|EPI_ISL_611407|20</td><td>····················</td><td>····················</td><td>····················</td><td>····················</td><td>····T···············</td><td>····················</td></tr>
<tr><td>O</td><td>427</td><td>hCoV-19/England/QEUH-A9F6FB/2020|EPI_ISL_611408|20</td><td>····················</td><td>····················</td><td>····················</td><td>····················</td><td>····················</td><td>····················</td></tr>
<tr><td>O</td><td>428</td><td>hCoV-19/England/QEUH-A9F35E/2020|EPI_ISL_611409|20</td><td>····················</td><td>····················</td><td>····················</td><td>····················</td><td>····················</td><td>····················</td></tr>
<tr><td>O</td><td>429</td><td>hCoV-19/England/QEUH-AA03B4/2020|EPI_ISL_611410|20</td><td>····················</td><td>····················</td><td>····················</td><td>····················</td><td>····················</td><td>····················</td></tr>
<tr><td>O</td><td>430</td><td>hCoV-19/England/QEUH-A9B729/2020|EPI_ISL_611411|20</td><td>····················</td><td>····················</td><td>····················</td><td>····················</td><td>····················</td><td>····················</td></tr>
</table>

| | | | | | | | | |
|---|---|---|---|---|---|---|---|---|
| O | 431 hCoV-19/England/QEUH-A9F50D/2020\|EPI_ISL_611412\|20 | ···················· | ···················· | ···················· | ···················· | ···················· | ···················· | |
| O | 432 hCoV-19/England/QEUH-A9F619/2020\|EPI_ISL_611413\|20 | ···················· | ···················· | ···················· | ···················· | ···················· | ···················· | |
| O | 433 hCoV-19/England/QEUH-AA035A/2020\|EPI_ISL_611414\|20 | ···················· | ···················· | ···················· | ···················· | ···················· | ···················· | |
| O | 434 hCoV-19/England/QEUH-A9ED7F/2020\|EPI_ISL_611415\|20 | ···················· | ···················· | ···················· | ···················· | ···················· | ···················· | |
| O | 435 hCoV-19/England/QEUH-AA024E/2020\|EPI_ISL_611416\|20 | ···················· | ···················· | ···················· | ···················· | ···················· | ···················· | |
| O | 436 hCoV-19/England/QEUH-A9EA4B/2020\|EPI_ISL_611417\|20 | ···················· | ···················· | ···················· | ···················· | ···················· | ···················· | |
| O | 437 hCoV-19/England/QEUH-A9EF4C/2020\|EPI_ISL_611418\|20 | ···················· | ···················· | ···················· | ···················· | ···················· | ···················· | |
| O | 438 hCoV-19/England/QEUH-A9AE9E/2020\|EPI_ISL_611419\|20 | ···················· | ···················· | ···················· | ···················· | ···················· | ···················· | |
| O | 439 hCoV-19/England/QEUH-A9B6FF/2020\|EPI_ISL_611420\|20 | ···················· | ···················· | ···················· | ···················· | ···················· | ···················· | |
| O | 440 hCoV-19/England/QEUH-A9FDE7/2020\|EPI_ISL_611421\|20 | ···················· | ···················· | ···················· | ···················· | ···················· | ···················· | |
| O | 441 hCoV-19/England/QEUH-A9F628/2020\|EPI_ISL_611422\|20 | ···················· | ···················· | ···················· | ···················· | ···················· | ···················· | |
| O | 442 hCoV-19/England/QEUH-A9EC36/2020\|EPI_ISL_611423\|20 | ···················· | ···················· | ···················· | ···················· | ···················· | ···················· | |
| O | 443 hCoV-19/England/QEUH-A9FB29/2020\|EPI_ISL_611424\|20 | ···················· | ···················· | ···················· | ···················· | ···················· | ···················· | |
| O | 444 hCoV-19/England/QEUH-A9F5C1/2020\|EPI_ISL_611425\|20 | ···················· | ···················· | ···················· | ···················· | ···················· | ···················· | |
| O | 445 hCoV-19/England/QEUH-A9F488/2020\|EPI_ISL_611426\|20 | ···················· | ···················· | ···················· | ···················· | ···················· | ···················· | |
| O | 446 hCoV-19/England/QEUH-AA028A/2020\|EPI_ISL_611427\|20 | ···················· | ···················· | ···················· | ···················· | ···················· | ···················· | |
| O | 447 hCoV-19/England/QEUH-A9ED42/2020\|EPI_ISL_611428\|20 | ···················· | ···················· | ···················· | ···················· | ···················· | ···················· | |
| O | 448 hCoV-19/England/QEUH-A9B695/2020\|EPI_ISL_611429\|20 | ···················· | ···················· | ···················· | ···················· | ···················· | ···················· | |
| O | 449 hCoV-19/England/QEUH-A9F45B/2020\|EPI_ISL_611430\|20 | ···················· | ···················· | ···················· | ···················· | ···················· | ···················· | |
| O | 450 hCoV-19/England/QEUH-AA03A5/2020\|EPI_ISL_611431\|20 | ···················· | ···················· | ···················· | ···················· | ···················· | ···················· | |
| O | 451 hCoV-19/England/QEUH-A9E89C/2020\|EPI_ISL_611432\|20 | ···················· | ···················· | ···················· | ···················· | ···················· | ···················· | |
| O | 452 hCoV-19/England/QEUH-AA02C6/2020\|EPI_ISL_611434\|20 | ···················· | ···················· | ···················· | ···················· | ···················· | ···················· | |
| O | 453 hCoV-19/England/QEUH-AA02B7/2020\|EPI_ISL_611435\|20 | ···················· | ···················· | ···················· | ···················· | ···················· | ···················· | |
| O | 454 hCoV-19/England/QEUH-A9F02A/2020\|EPI_ISL_611436\|20 | ···················· | ···················· | ···················· | ···················· | ···················· | ···················· | |
| O | 455 hCoV-19/England/QEUH-A9F5FE/2020\|EPI_ISL_611437\|20 | ···················· | ···················· | ···················· | ···················· | ···················· | ···················· | |
| O | 456 hCoV-19/England/QEUH-A9F576/2020\|EPI_ISL_611438\|20 | ···················· | ···················· | ···················· | ···················· | ···················· | ···················· | |
| O | 457 hCoV-19/England/QEUH-A9F585/2020\|EPI_ISL_611439\|20 | ···················· | ···················· | ···················· | ···················· | ···················· | ···················· | |
| O | 458 hCoV-19/England/QEUH-A9F127/2020\|EPI_ISL_611440\|20 | ···················· | ···················· | ···················· | ···················· | ···················· | ···················· | |
| O | 459 hCoV-19/England/QEUH-A9FE3F/2020\|EPI_ISL_611441\|20 | ···················· | ···················· | ···················· | ···················· | ···················· | ···················· | |
| O | 460 hCoV-19/England/QEUH-A9FE02/2020\|EPI_ISL_611442\|20 | ···················· | ···················· | ···················· | ···················· | ···················· | ···················· | |
| O | 461 hCoV-19/England/QEUH-A9EF3D/2020\|EPI_ISL_611444\|20 | ···················· | ·A·················· | ···················· | ···················· | ···················· | ··A················· | |
| O | 462 hCoV-19/England/QEUH-A9E9B7/2020\|EPI_ISL_611445\|20 | ···················· | ···················· | ···················· | ···················· | ·····T·············· | ···················· | |
| O | 463 hCoV-19/England/QEUH-A9F39A/2020\|EPI_ISL_611447\|20 | ···················· | ···················· | ···················· | ···················· | ···················· | ···················· | |
| O | 464 hCoV-19/England/QEUH-A9FBFC/2020\|EPI_ISL_611480\|20 | ···················· | ·A·················· | ···················· | ···················· | ···················· | ··A················· | |
| O | 465 hCoV-19/England/QEUH-A9FBB0/2020\|EPI_ISL_611481\|20 | ···················· | ···················· | ···················· | ···················· | ···················· | ···················· | |
| O | 466 hCoV-19/England/QEUH-AA01F6/2020\|EPI_ISL_611482\|20 | ···················· | ···················· | ···················· | ···················· | ···················· | ···················· | |
| O | 467 hCoV-19/England/QEUH-A9F691/2020\|EPI_ISL_611483\|20 | ···················· | ·A·················· | ···················· | ···················· | ···················· | ··A················· | |
| O | 468 hCoV-19/England/QEUH-AA01BA/2020\|EPI_ISL_611484\|20 | ···················· | ···················· | ···················· | ···················· | ···················· | ···················· | |
| O | 469 hCoV-19/Wales/PHWC-47F879/2020\|EPI_ISL_612735\|2020 | ···················· | ···················· | ···················· | ···················· | ···················· | ···················· | |
| O | 470 hCoV-19/Wales/PHWC-47F888/2020\|EPI_ISL_612736\|2020 | ···················· | ···················· | ···················· | ···················· | ···················· | ···················· | |
| O | 471 hCoV-19/Wales/PHWC-47F897/2020\|EPI_ISL_612737\|2020 | ···················· | ···················· | ···················· | ···················· | ···················· | ···················· | |
| O | 472 hCoV-19/Wales/PHWC-47F8A6/2020\|EPI_ISL_612738\|2020 | ···················· | ···················· | ···················· | ···················· | ···················· | ···················· | |
| O | 473 hCoV-19/Wales/PHWC-47F8B5/2020\|EPI_ISL_612739\|2020 | ···················· | ···················· | ···················· | ···················· | ···················· | ···················· | |
| O | 474 hCoV-19/Wales/PHWC-47F8C4/2020\|EPI_ISL_612740\|2020 | ···················· | ···················· | ···················· | ···················· | NNN················· | ···················· | 1 |
| O | 475 hCoV-19/Wales/PHWC-47F8D3/2020\|EPI_ISL_612741\|2020 | ···················· | ···················· | ···················· | ···················· | ···················· | ···················· | |
| O | 476 hCoV-19/Wales/PHWC-47F8F1/2020\|EPI_ISL_612742\|2020 | ···················· | ···················· | ···················· | ···················· | ···················· | ···················· | |
| O | 477 hCoV-19/Wales/PHWC-47F90D/2020\|EPI_ISL_612743\|2020 | ···················· | ···················· | ···················· | ···················· | ···················· | ···················· | |
| O | 478 hCoV-19/Wales/PHWC-47F91C/2020\|EPI_ISL_612744\|2020 | ···················· | ···················· | ···················· | ···················· | ···················· | ···················· | |
| O | 479 hCoV-19/Wales/PHWC-47F92B/2020\|EPI_ISL_612745\|2020 | ···················· | ···················· | ···················· | ···················· | ···················· | ···················· | |
| O | 480 hCoV-19/Wales/PHWC-47F949/2020\|EPI_ISL_612746\|2020 | ···················· | ···················· | ···················· | ···················· | ···················· | ···················· | |
| O | 481 hCoV-19/Wales/PHWC-47F958/2020\|EPI_ISL_612747\|2020 | ···················· | ···················· | ···················· | ···················· | ···················· | ···················· | |
| O | 482 hCoV-19/Wales/PHWC-47F967/2020\|EPI_ISL_612748\|2020 | ···················· | ···················· | ···················· | ···················· | ···················· | ···················· | |
| O | 483 hCoV-19/Wales/PHWC-47F976/2020\|EPI_ISL_612749\|2020 | ···················· | ···················· | ···················· | ···················· | ···················· | ···················· | |
| O | 484 hCoV-19/Wales/PHWC-47F985/2020\|EPI_ISL_612750\|2020 | ···················· | ···················· | ···················· | ···················· | ···················· | ···················· | |
| O | 485 hCoV-19/Wales/PHWC-47F994/2020\|EPI_ISL_612751\|2020 | ···················· | ···················· | ···················· | ···················· | ···················· | ···················· | |
| O | 486 hCoV-19/Wales/PHWC-47F9A3/2020\|EPI_ISL_612752\|2020 | ···················· | ···················· | ···················· | ···················· | ···················· | ···················· | |
| O | 487 hCoV-19/Wales/PHWC-47F9B2/2020\|EPI_ISL_612753\|2020 | ···················· | ···················· | ···················· | ···················· | ···················· | ···················· | |
| O | 488 hCoV-19/Wales/PHWC-47F9C1/2020\|EPI_ISL_612754\|2020 | ···················· | ···················· | ···················· | ···················· | ···················· | ···················· | |
| O | 489 hCoV-19/Wales/PHWC-47F9D0/2020\|EPI_ISL_612755\|2020 | ···················· | ···················· | ···················· | ···················· | ···················· | ···················· | |
| O | 490 hCoV-19/Wales/PHWC-47F9EF/2020\|EPI_ISL_612756\|2020 | ···················· | ···················· | ···················· | ···················· | ···················· | ···················· | |
| O | 491 hCoV-19/Wales/PHWC-47F9FE/2020\|EPI_ISL_612757\|2020 | ···················· | ···················· | ···················· | ···················· | ···················· | ···················· | |
| O | 492 hCoV-19/Wales/PHWC-47FA0A/2020\|EPI_ISL_612758\|2020 | ···················· | ···················· | ···················· | ···················· | ···················· | ···················· | |
| O | 493 hCoV-19/Wales/PHWC-47FA19/2020\|EPI_ISL_612759\|2020 | ···················· | ···················· | ···················· | ···················· | ···················· | ···················· | |
| O | 494 hCoV-19/Wales/PHWC-47FA28/2020\|EPI_ISL_612760\|2020 | ···················· | ···················· | ···················· | ···················· | ···················· | ···················· | |
| O | 495 hCoV-19/Wales/PHWC-47FA37/2020\|EPI_ISL_612761\|2020 | ···················· | ···················· | ···················· | ···················· | ···················· | ···················· | |
| O | 496 hCoV-19/Wales/PHWC-47FA46/2020\|EPI_ISL_612762\|2020 | ···················· | ···················· | ···················· | ···················· | ···················· | ···················· | |
| O | 497 hCoV-19/Wales/PHWC-47FA55/2020\|EPI_ISL_612763\|2020 | ···················· | ···················· | ···················· | ···················· | ···················· | ···················· | |
| O | 498 hCoV-19/Wales/PHWC-47FA64/2020\|EPI_ISL_612764\|2020 | ···················· | ···················· | ···················· | ···················· | ···················· | ···················· | |
| O | 500 hCoV-19/Wales/PHWC-47FA91/2020\|EPI_ISL_612766\|2020 | ···················· | ···················· | ···················· | ···················· | ···················· | ···················· | |
| O | 501 hCoV-19/Wales/PHWC-47FAA0/2020\|EPI_ISL_612767\|2020 | ···················· | ···················· | ···················· | ···················· | ···················· | ···················· | |
| O | 502 hCoV-19/Wales/PHWC-47FABF/2020\|EPI_ISL_612768\|2020 | ···················· | ···················· | ···················· | ···················· | ···················· | ···················· | |
| O | 503 hCoV-19/Wales/PHWC-47FACE/2020\|EPI_ISL_612769\|2020 | ···················· | ···················· | ···················· | ···················· | ···················· | ···················· | |
| O | 504 hCoV-19/Wales/PHWC-47FADD/2020\|EPI_ISL_612770\|2020 | ···················· | ···················· | ···················· | ···················· | NNN················· | ···················· | 1 |
| O | 505 hCoV-19/Wales/PHWC-47FAFB/2020\|EPI_ISL_612771\|2020 | ···················· | ···················· | ···················· | ···················· | ···················· | ···················· | |
| O | 506 hCoV-19/Wales/PHWC-47FB07/2020\|EPI_ISL_612772\|2020 | ···················· | ···················· | ···················· | ···················· | ···················· | ···················· | |
| O | 507 hCoV-19/Wales/PHWC-47FB16/2020\|EPI_ISL_612773\|2020 | ···················· | ···················· | ···················· | ···················· | ···················· | ···················· | |
| O | 508 hCoV-19/Wales/PHWC-47FB25/2020\|EPI_ISL_612774\|2020 | ···················· | ···················· | ···················· | ···················· | ···················· | ···················· | |
| O | 510 hCoV-19/Wales/PHWC-47FB52/2020\|EPI_ISL_612776\|2020 | ···················· | ···················· | ···················· | ···················· | ···················· | ···················· | |
| O | 511 hCoV-19/Wales/PHWC-47FB61/2020\|EPI_ISL_612777\|2020 | ···················· | ···················· | ···················· | ···················· | ···················· | ···················· | |
| O | 512 hCoV-19/Wales/PHWC-47FB70/2020\|EPI_ISL_612778\|2020 | ···················· | ···················· | ···················· | ···················· | ···················· | ···················· | |

| | | | | | | | | |
|---|---|---|---|---|---|---|---|---|
| O | 513 | hCoV-19/England/MILK-A7ACB8/2020\|EPI_ISL_606800\|20 | ···················· | ···················· | ····················· | ···················· | ···················· | ···················· |
| O | 514 | hCoV-19/England/MILK-A7CE0B/2020\|EPI_ISL_606801\|20 | ···················· | ···················· | ····················· | ···················· | ···················· | ···················· |
| O | 515 | hCoV-19/England/MILK-A79B70/2020\|EPI_ISL_606803\|20 | ···················· | ···················· | ····················· | ···················· | ···················· | ···················· |
| O | 516 | hCoV-19/England/MILK-A7A69C/2020\|EPI_ISL_606804\|20 | ···················· | ···················· | ····················· | ···················· | ···················· | ···················· |
| O | 517 | hCoV-19/England/MILK-A7ADD3/2020\|EPI_ISL_606805\|20 | ···················· | ···················· | ····················· | ···················· | ···················· | ···················· |
| O | 518 | hCoV-19/England/MILK-A7CFBD/2020\|EPI_ISL_606807\|20 | ···················· | ···················· | ····················· | ···················· | ···················· | ···················· |
| O | 519 | hCoV-19/England/MILK-A7A395/2020\|EPI_ISL_606808\|20 | ···················· | ···················· | ····················· | ···················· | ···················· | ···················· |
| O | 520 | hCoV-19/England/MILK-A7ACE5/2020\|EPI_ISL_606809\|20 | ···················· | ···················· | ····················· | ···················· | ·····T·············· | ···················· |
| O | 521 | hCoV-19/England/MILK-A7A016/2020\|EPI_ISL_606810\|20 | ···················· | ···················· | ····················· | ···················· | ···················· | ···················· |
| O | 522 | hCoV-19/England/MILK-A797A9/2020\|EPI_ISL_606811\|20 | ···················· | ···················· | ····················· | ···················· | ···················· | ···················· |
| O | 523 | hCoV-19/England/MILK-A7A007/2020\|EPI_ISL_606812\|20 | ···················· | ···················· | ····················· | ···················· | ···················· | ···················· |
| O | 524 | hCoV-19/England/MILK-A7D532/2020\|EPI_ISL_606813\|20 | ···················· | ···················· | ····················· | ···················· | ···················· | ···················· |
| O | 525 | hCoV-19/England/MILK-A79D98/2020\|EPI_ISL_606814\|20 | ···················· | ···················· | ····················· | ···················· | ···················· | ···················· |
| O | 526 | hCoV-19/England/MILK-A79AFB/2020\|EPI_ISL_606815\|20 | ···················· | ···················· | ····················· | ···················· | ···················· | ···················· |
| O | 527 | hCoV-19/England/MILK-A7975E/2020\|EPI_ISL_606816\|20 | ···················· | ···················· | ····················· | ···················· | ···················· | ···················· |
| O | 528 | hCoV-19/England/MILK-A796D9/2020\|EPI_ISL_606860\|20 | ···················· | ···················· | ····················· | ···················· | ·····T·············· | ···················· |
| O | 529 | hCoV-19/England/MILK-A799C1/2020\|EPI_ISL_606864\|20 | ···················· | ···················· | ····················· | ···················· | ···················· | ···················· |
| O | 530 | hCoV-19/England/MILK-A7CF26/2020\|EPI_ISL_606865\|20 | ···················· | ···················· | ····················· | ···················· | ···················· | ···················· |
| O | 531 | hCoV-19/England/MILK-A7A40B/2020\|EPI_ISL_606869\|20 | ···················· | ···················· | ····················· | ···················· | ···················· | ···················· |
| O | 532 | hCoV-19/England/MILK-A79DF2/2020\|EPI_ISL_606870\|20 | ···················· | ···················· | ····················· | ···················· | ···················· | ···················· |
| O | 533 | hCoV-19/England/MILK-A79651/2020\|EPI_ISL_606871\|20 | ···················· | ·A·················· | ····················· | ···················· | ···················· | ···A················ |
| O | 534 | hCoV-19/England/MILK-A79C7D/2020\|EPI_ISL_606872\|20 | ···················· | ···················· | ····················· | ···················· | ···················· | ···················· |
| O | 535 | hCoV-19/England/MILK-A79ACE/2020\|EPI_ISL_606873\|20 | ···················· | ···················· | ····················· | ···················· | ···················· | ···················· |
| O | 536 | hCoV-19/England/MILK-A7E8CF/2020\|EPI_ISL_606874\|20 | ···················· | ···················· | ····················· | ···················· | ···················· | ···················· |
| O | 537 | hCoV-19/England/MILK-A796AC/2020\|EPI_ISL_606876\|20 | ···················· | ···················· | ····················· | ···················· | ···················· | ···················· |
| O | 538 | hCoV-19/England/MILK-A7A4CF/2020\|EPI_ISL_606877\|20 | ···················· | ···················· | ····················· | ···················· | ···················· | ···················· |
| O | 539 | hCoV-19/England/MILK-A79DB6/2020\|EPI_ISL_606881\|20 | ···················· | ···················· | ····················· | ···················· | ···················· | ···················· |
| O | 540 | hCoV-19/England/MILK-A79A91/2020\|EPI_ISL_606882\|20 | ···················· | ···················· | ····················· | ···················· | ···················· | ···················· |
| O | 541 | hCoV-19/England/MILK-A7A517/2020\|EPI_ISL_606883\|20 | ···················· | ···················· | ····················· | ···················· | ···················· | ···················· |
| O | 542 | hCoV-19/England/MILK-A7A19B/2020\|EPI_ISL_606884\|20 | ···················· | ···················· | ····················· | ···················· | ···················· | ···················· |
| O | 543 | hCoV-19/England/MILK-A79E3B/2020\|EPI_ISL_606885\|20 | ···················· | ···················· | ····················· | ···················· | ···················· | ···················· |
| O | 544 | hCoV-19/England/MILK-A7A605/2020\|EPI_ISL_606886\|20 | ···················· | ···················· | ····················· | ···················· | ···················· | ···················· |
| O | 545 | hCoV-19/England/MILK-A799D0/2020\|EPI_ISL_606888\|20 | ···················· | ···················· | ····················· | ···················· | ···················· | ···················· |
| O | 546 | hCoV-19/England/MILK-A796F7/2020\|EPI_ISL_606889\|20 | ···················· | ···················· | ····················· | ···················· | ···················· | ···················· |
| O | 547 | hCoV-19/England/MILK-A79A64/2020\|EPI_ISL_606890\|20 | ···················· | ·A·················· | ····················· | ···················· | ···················· | ···A················ |
| O | 548 | hCoV-19/England/MILK-A79703/2020\|EPI_ISL_606891\|20 | ···················· | ···················· | ····················· | ···················· | ···················· | ···················· |
| O | 549 | hCoV-19/England/MILK-A795AF/2020\|EPI_ISL_606893\|20 | ···················· | ·A·················· | ····················· | ···················· | ···················· | ···················· |
| O | 550 | hCoV-19/England/MILK-A7CCE3/2020\|EPI_ISL_606895\|20 | ···················· | ·A·················· | ····················· | ···················· | ···················· | ···A················ |
| O | 551 | hCoV-19/England/MILK-A7A122/2020\|EPI_ISL_606896\|20 | ···················· | ···················· | ····················· | ···················· | ···················· | ···················· |
| O | 552 | hCoV-19/England/MILK-A7A0DA/2020\|EPI_ISL_606898\|20 | ···················· | ···················· | ····················· | ···················· | ···················· | ···················· |
| O | 553 | hCoV-19/England/MILK-A79D10/2020\|EPI_ISL_606899\|20 | ···················· | ···················· | ····················· | ···················· | ···················· | ···················· |
| O | 554 | hCoV-19/England/ALDP-A690B0/2020\|EPI_ISL_609010\|20 | ···················· | ···················· | ····················· | ···················· | ···················· | ···················· |
| O | 555 | hCoV-19/England/ALDP-A693E4/2020\|EPI_ISL_609011\|20 | ···················· | ···················· | ····················· | ···················· | ···················· | ···················· |
| O | 556 | hCoV-19/England/ALDP-A692E7/2020\|EPI_ISL_609012\|20 | ···················· | ···················· | ····················· | ···················· | ···················· | ···················· |
| O | 557 | hCoV-19/England/ALDP-A689D4/2020\|EPI_ISL_609013\|20 | ···················· | ···················· | ····················· | ···················· | ···················· | ···················· |
| O | 558 | hCoV-19/England/CAMC-A6463B/2020\|EPI_ISL_609014\|20 | ···················· | ···················· | ····················· | ···················· | ···················· | ···················· |
| O | 559 | hCoV-19/England/ALDP-A694C3/2020\|EPI_ISL_609016\|20 | ···················· | ···················· | ····················· | ···················· | ···················· | ···················· |
| O | 560 | hCoV-19/England/MILK-A6C071/2020\|EPI_ISL_609017\|20 | ···················· | ···················· | ····················· | ···················· | ···················· | ···················· |
| O | 561 | hCoV-19/England/ALDP-A67D06/2020\|EPI_ISL_609019\|20 | ···················· | ···················· | ····················· | ···················· | ···················· | ···················· |
| O | 562 | hCoV-19/England/ALDP-A69645/2020\|EPI_ISL_609020\|20 | ···················· | ···················· | ····················· | ···················· | ···················· | ···················· |
| O | 563 | hCoV-19/England/ALDP-A69214/2020\|EPI_ISL_609022\|20 | ···················· | ···················· | ····················· | ···················· | ···················· | ···················· |
| O | 564 | hCoV-19/England/CAMC-A64A20/2020\|EPI_ISL_609023\|20 | ···················· | ···················· | ····················· | ···················· | ···················· | ···················· |
| O | 565 | hCoV-19/England/CAMC-A647DE/2020\|EPI_ISL_609025\|20 | ···················· | ···················· | ····················· | ···················· | ···················· | ···················· |
| O | 566 | hCoV-19/England/ALDP-A682D9/2020\|EPI_ISL_609028\|20 | ···················· | ···················· | ····················· | ···················· | ···················· | ···················· |
| O | 567 | hCoV-19/England/MILK-A6BE1E/2020\|EPI_ISL_609029\|20 | ···················· | ···················· | ····················· | ···················· | ···················· | ···················· |
| O | 568 | hCoV-19/England/MILK-A6BE0F/2020\|EPI_ISL_609030\|20 | ···················· | ···················· | ····················· | ···················· | ···················· | ···················· |
| O | 569 | hCoV-19/England/ALDP-A6802A/2020\|EPI_ISL_609031\|20 | ···················· | ···················· | ····················· | ···················· | ···················· | ···················· |
| O | 570 | hCoV-19/England/ALDP-A694B4/2020\|EPI_ISL_609032\|20 | ···················· | ···················· | ····················· | ···················· | ···················· | ···················· |
| O | 571 | hCoV-19/England/CAMC-A646A4/2020\|EPI_ISL_609033\|20 | ···················· | ···················· | ····················· | ···················· | ···················· | ···················· |
| O | 572 | hCoV-19/England/ALDP-A69108/2020\|EPI_ISL_609034\|20 | ···················· | ···················· | ····················· | ···················· | ···················· | ···················· |
| O | 573 | hCoV-19/England/ALDP-A6936C/2020\|EPI_ISL_609035\|20 | ···················· | ···················· | ····················· | ···················· | ···················· | ···················· |
| O | 574 | hCoV-19/England/MILK-A6C141/2020\|EPI_ISL_609036\|20 | ···················· | ·········T·········· | ····················· | ···················· | ···················· | ···················· |
| O | 575 | hCoV-19/England/CAMC-A64C93/2020\|EPI_ISL_609037\|20 | ···················· | ···················· | ····················· | ···················· | ···················· | ···················· |
| O | 576 | hCoV-19/England/MILK-A6BDE4/2020\|EPI_ISL_609038\|20 | ···················· | ···················· | ····················· | ···················· | ···················· | ···················· |
| O | 577 | hCoV-19/England/ALDP-A68048/2020\|EPI_ISL_609039\|20 | ···················· | ···················· | ····················· | ···················· | ···················· | ···················· |
| O | 578 | hCoV-19/England/ALDP-A69469/2020\|EPI_ISL_609040\|20 | ···················· | ···················· | ····················· | ···················· | ···················· | ···················· |
| O | 579 | hCoV-19/England/ALDP-A69232/2020\|EPI_ISL_609041\|20 | ···················· | ·A·················· | ····················· | ···················· | ···················· | ···A················ |
| O | 580 | hCoV-19/England/MILK-A6BE2D/2020\|EPI_ISL_609042\|20 | ···················· | ···················· | ····················· | ···················· | ···················· | ···················· |
| O | 581 | hCoV-19/England/ALDP-A68109/2020\|EPI_ISL_609043\|20 | ···················· | ···················· | ····················· | ···················· | ···················· | ···················· |
| O | 582 | hCoV-19/England/CAMC-A648BD/2020\|EPI_ISL_609045\|20 | ···················· | ···················· | ····················· | ···················· | ···················· | ···················· |
| O | 583 | hCoV-19/England/MILK-A6BD4E/2020\|EPI_ISL_609046\|20 | ···················· | ···················· | ····················· | ···················· | ···················· | ···················· |
| O | 584 | hCoV-19/England/ALDP-A68093/2020\|EPI_ISL_609047\|20 | ···················· | ···················· | ····················· | ···················· | ···················· | ···················· |
| O | 585 | hCoV-19/England/MILK-A6C16F/2020\|EPI_ISL_609049\|20 | ···················· | ···················· | ····················· | ···················· | ···················· | ···················· |
| O | 586 | hCoV-19/England/ALDP-A68B29/2020\|EPI_ISL_609070\|20 | ···················· | ···················· | ····················· | ···················· | ···················· | ···················· |
| O | 587 | hCoV-19/England/MILK-A6BD02/2020\|EPI_ISL_609071\|20 | ···················· | ···················· | ····················· | ···················· | ···················· | ···················· |
| O | 588 | hCoV-19/England/CAMC-A64808/2020\|EPI_ISL_609072\|20 | ···················· | ···················· | ····················· | ···················· | ···················· | ···················· |
| O | 589 | hCoV-19/England/ALDP-A67FF1/2020\|EPI_ISL_609074\|20 | ···················· | ···················· | ····················· | ···················· | ···················· | ···················· |
| O | 590 | hCoV-19/England/ALDP-A680EE/2020\|EPI_ISL_609078\|20 | ···················· | ···················· | ····················· | ···················· | ···················· | ···················· |
| O | 591 | hCoV-19/England/ALDP-A690ED/2020\|EPI_ISL_609079\|20 | ···················· | ·A·················· | ····················· | ···················· | ···················· | ···A················ |
| O | 592 | hCoV-19/England/ALDP-A6897A/2020\|EPI_ISL_609080\|20 | ···················· | ···················· | ····················· | ···················· | ···················· | ···················· |

O 593 hCoV-19/England/CAMC-A64A11/2020|EPI_ISL_609081|20 .................... .................... .................... .................... .................... ....................

O 594 hCoV-19/England/ALDP-A69047/2020|EPI_ISL_609083|20 .................... .................... .................... .................... .................... ....................

O 595 hCoV-19/England/MILK-A6BDD5/2020|EPI_ISL_609084|20 .................... .................... .................... .................... .................... ....................

O 596 hCoV-19/England/ALDP-A69302/2020|EPI_ISL_609085|20 .................... .................... .................... .................... .................... ....................

O 597 hCoV-19/England/CAMC-A645A7/2020|EPI_ISL_609086|20 .................... .................... .................... .................... .................... ....................

O 598 hCoV-19/England/ALDP-A69663/2020|EPI_ISL_609087|20 .................... .................... .................... .................... .................... ....................

O 599 hCoV-19/England/CAMC-A648F9/2020|EPI_ISL_609088|20 .................... .................... .................... .................... .................... ....................

O 600 hCoV-19/England/ALDP-A69636/2020|EPI_ISL_609089|20 .................... .................... .................... .................... .................... ....................

O 601 hCoV-19/USA/MI-UM-10036413655/2020|EPI_ISL_610060| .................... .................... .................... .................... .................... ....................

O 604 hCoV-19/USA/MI-UM-10036400561/2020|EPI_ISL_610063| .................... .................... .................... .................... .................... ....................

O 606 hCoV-19/USA/MI-UM-10036386923/2020|EPI_ISL_610069| .................... .................... .................... .................... .................... ....................

O 609 hCoV-19/USA/MI-UM-10036409513/2020|EPI_ISL_610072| .................... .................... .................... .................... .................... ....................

O 610 hCoV-19/USA/MI-UM-10036396949/2020|EPI_ISL_610073| .................... .................... .................... .................... .................... ....................

O 611 hCoV-19/USA/MI-UM-10036428041/2020|EPI_ISL_610074| .................... .................... .................... .................... .................... ....................

O 612 hCoV-19/USA/MI-UM-10036404422/2020|EPI_ISL_610075| .................... .................... .................... .................... .................... ....................

O 613 hCoV-19/USA/MI-UM-10036391877/2020|EPI_ISL_610076| .................... .A.................. .................... .................... .................... ....................

O 614 hCoV-19/USA/MI-UM-10036409812/2020|EPI_ISL_610078| .................... .................... .................... .................... .................... ....................

O 615 hCoV-19/USA/MI-UM-10036428187/2020|EPI_ISL_610079| .................... .................... .................... .................... .................... ....................

O 616 hCoV-19/England/QEUH-A9B738/2020|EPI_ISL_611327|20 .................... .................... .................... .................... .................... ....................

O 617 hCoV-19/England/QEUH-A9FB74/2020|EPI_ISL_611328|20 .................... .................... .................... .................... .................... ....................

O 618 hCoV-19/England/QEUH-A9EF00/2020|EPI_ISL_611329|20 .................... .................... .................... .................... .................... ....................

O 619 hCoV-19/England/QEUH-A9F43D/2020|EPI_ISL_611330|20 .................... .................... .................... .................... .................... ....................

O 620 hCoV-19/England/QEUH-AA040C/2020|EPI_ISL_611331|20 .................... .................... .................... .................... .................... ....................

O 621 hCoV-19/England/QEUH-A9F41F/2020|EPI_ISL_611332|20 .................... .................... .................... .................... .................... ....................

O 622 hCoV-19/England/QEUH-A9FDD8/2020|EPI_ISL_611333|20 .................... .................... .................... .................... .................... ....................

O 623 hCoV-19/England/QEUH-AA03D2/2020|EPI_ISL_611334|20 .................... .................... .................... .................... .................... ...A................

O 624 hCoV-19/England/QEUH-A9E9D5/2020|EPI_ISL_611335|20 .................... .A.................. .................... .................... .................... ....................

O 625 hCoV-19/England/QEUH-A9F0C0/2020|EPI_ISL_611336|20 .................... .................... .................... .................... .................... ....................

O 626 hCoV-19/England/QEUH-A9ED15/2020|EPI_ISL_611337|20 .................... .................... .................... .................... .................... ....................

O 627 hCoV-19/England/QEUH-A9F4D3/2020|EPI_ISL_611338|20 .................... .................... .................... .................... .................... ....................

O 628 hCoV-19/England/QEUH-A9FE99/2020|EPI_ISL_611339|20 .................... .................... .................... .................... .................... ....................

O 629 hCoV-19/England/QEUH-AA033C/2020|EPI_ISL_611340|20 .................... .................... .................... .................... .................... ....................

O 630 hCoV-19/England/QEUH-A9EE9A/2020|EPI_ISL_611341|20 .................... .................... .................... .................... .................... ....................

O 631 hCoV-19/England/QEUH-A9F3F4/2020|EPI_ISL_611342|20 .................... .................... .................... .................... .................... ....................

O 632 hCoV-19/England/QEUH-A9B774/2020|EPI_ISL_611343|20 .................... .................... .................... .................... .................... ....................

O 633 hCoV-19/England/QEUH-AA0590/2020|EPI_ISL_611344|20 .................... .................... .................... .................... .................... ....................

O 634 hCoV-19/England/QEUH-A9B70B/2020|EPI_ISL_611345|20 .................... .................... .................... .................... .................... ....................

O 635 hCoV-19/England/QEUH-AA0008/2020|EPI_ISL_611348|20 .................... .................... .................... .................... .................... ....................

O 636 hCoV-19/England/QEUH-A9FB1A/2020|EPI_ISL_611349|20 .................... .................... .................... .................... .................... ....................

O 637 hCoV-19/England/QEUH-AA0545/2020|EPI_ISL_611350|20 .................... .................... .................... .................... .................... ....................

O 638 hCoV-19/England/QEUH-A9EC63/2020|EPI_ISL_611351|20 .................... .A.................. .................... .................... .................... ....................

O 639 hCoV-19/England/QEUH-A9F400/2020|EPI_ISL_611352|20 .................... .................... .................... .................... .................... ....................

O 640 hCoV-19/England/QEUH-A9F6A0/2020|EPI_ISL_611353|20 .................... .................... .................... .................... .................... ....................

O 641 hCoV-19/England/QEUH-AA0369/2020|EPI_ISL_611354|20 .................... .................... .................... .................... .................... ....................

O 642 hCoV-19/England/QEUH-A9FBDE/2020|EPI_ISL_611355|20 .................... .................... .................... .................... .................... ....................

O 643 hCoV-19/England/QEUH-A9F01B/2020|EPI_ISL_611356|20 .................... .................... .................... .................... .................... ....................

O 644 hCoV-19/England/QEUH-AA032D/2020|EPI_ISL_611357|20 .................... .................... .................... .................... .................... ....................

O 645 hCoV-19/England/QEUH-A9FB56/2020|EPI_ISL_611358|20 .................... .................... .................... .................... ......T............. ....................

O 646 hCoV-19/England/QEUH-A9F36D/2020|EPI_ISL_611359|20 .................... .................... .................... .................... .................... ....................

O 647 hCoV-19/England/QEUH-A9F6BF/2020|EPI_ISL_611360|20 .................... .................... .................... .................... .................... ....................

O 648 hCoV-19/England/QEUH-A9F34F/2020|EPI_ISL_611361|20 .................... .................... .................... .................... .................... ....................

O 649 hCoV-19/England/QEUH-A9B6A4/2020|EPI_ISL_611363|20 .................... .................... .................... .................... .................... ....................

O 650 hCoV-19/England/QEUH-A9F2CA/2020|EPI_ISL_611364|20 .................... .................... .................... .................... .................... ....................

O 651 hCoV-19/England/QEUH-A9B61D/2020|EPI_ISL_611365|20 .................... .................... .................... .................... .................... ....................

O 652 hCoV-19/England/QEUH-A9FE20/2020|EPI_ISL_611366|20 .................... .................... .................... .................... .................... ...A................

O 653 hCoV-19/England/QEUH-A9F3C7/2020|EPI_ISL_611367|20 .................... .................... .................... .................... .................... ....................

O 654 hCoV-19/England/QEUH-A9F673/2020|EPI_ISL_611368|20 .................... .................... .................... .................... .................... ....................

O 655 hCoV-19/England/QEUH-A9F38B/2020|EPI_ISL_611390|20 .................... .................... .................... .................... .................... ....................

O 656 hCoV-19/England/QEUH-A9F46A/2020|EPI_ISL_611391|20 .................... .................... .................... .................... .................... ....................

O 657 hCoV-19/England/QEUH-A9EE5E/2020|EPI_ISL_611392|20 .................... .................... .................... .................... .................... ....................

O 658 hCoV-19/England/QEUH-A9F109/2020|EPI_ISL_611393|20 .................... .................... .................... .................... .................... ....................

O 659 hCoV-19/England/QEUH-AA01AB/2020|EPI_ISL_611394|20 .................... .................... .................... .................... .................... ....................

O 660 hCoV-19/England/QEUH-A9F52B/2020|EPI_ISL_611395|20 .................... .................... .................... .................... .................... ....................

O 661 hCoV-19/England/NOTT-1183CF/2020|EPI_ISL_612600|20 .................... .................... .................... .................... .................... ....................

O 662 hCoV-19/England/NOTT-1183FC/2020|EPI_ISL_612601|20 .................... .................... .................... .................... .................... ....................

O 663 hCoV-19/England/NOTT-118408/2020|EPI_ISL_612602|20 .................... .................... .................... .................... .................... ....................

O 664 hCoV-19/England/NOTT-118426/2020|EPI_ISL_612603|20 .................... .................... .................... .................... .................... ....................

O 665 hCoV-19/England/NOTT-118435/2020|EPI_ISL_612604|20 .................... .................... .................... .................... .................... ....................

O 666 hCoV-19/England/NOTT-118444/2020|EPI_ISL_612605|20 .................... .................... .................... .................... .................... ....................

O 667 hCoV-19/England/NOTT-118471/2020|EPI_ISL_612606|20 .................... .................... .................... .................... .................... ....................

O 668 hCoV-19/Wales/PHWC-47EF66/2020|EPI_ISL_612607|2020 .................... .................... .................... .................... .................... ....................

O 669 hCoV-19/Wales/PHWC-47EF75/2020|EPI_ISL_612608|2020 .................... .................... .................... .................... .................... ....................

O 670 hCoV-19/Wales/PHWC-47EF84/2020|EPI_ISL_612609|2020 .................... .................... .................... .................... .................... ....................

O 671 hCoV-19/Wales/PHWC-47EF93/2020|EPI_ISL_612610|2020 .................... .................... .................... .................... .................... ....................

O 672 hCoV-19/Wales/PHWC-47EFA2/2020|EPI_ISL_612611|2020 .................... .................... .................... .................... .................... ....................

O 673 hCoV-19/Wales/PHWC-47EFB1/2020|EPI_ISL_612612|2020 .................... .................... .................... .................... .................... ....................

O 674 hCoV-19/Wales/PHWC-47EFC0/2020|EPI_ISL_612613|2020 .................... .................... .................... .................... .................... ....................

O 675 hCoV-19/Wales/PHWC-47F044/2020|EPI_ISL_612620|2020 .................... .................... .................... .................... .................... ....................

O 676 hCoV-19/Wales/PHWC-47F053/2020|EPI_ISL_612621|2020 .................... .................... .................... .................... .................... ....................

O 677 hCoV-19/Wales/PHWC-47F071/2020|EPI_ISL_612623|2020 .................... .................... .................... .................... .................... ....................

O 678 hCoV-19/Wales/PHWC-47F080/2020|EPI_ISL_612624|2020 .................... .................... ..................... .................... .................... ....................
O 679 hCoV-19/Wales/PHWC-47F2D5/2020|EPI_ISL_612658|2020 .................... .................... ..................... .................... .................... ....................
O 680 hCoV-19/Wales/PHWC-47F2E4/2020|EPI_ISL_612659|2020 .................... .................... ..................... .................... .................... ....................
O 681 hCoV-19/Wales/PHWC-47F30F/2020|EPI_ISL_612660|2020 .................... .................... ..................... .................... .................... ....................
O 682 hCoV-19/Wales/PHWC-47F32D/2020|EPI_ISL_612661|2020 .................... .................... ..................... .................... .................... ....................
O 683 hCoV-19/Wales/PHWC-47F33C/2020|EPI_ISL_612662|2020 .................... .................... ..................... .................... .................... ....................
O 684 hCoV-19/Wales/PHWC-47F34B/2020|EPI_ISL_612663|2020 .................... .................... ..................... .................... .................... ....................
O 685 hCoV-19/Wales/PHWC-47F35A/2020|EPI_ISL_612664|2020 .................... .................... ..................... .................... .................... ....................
O 686 hCoV-19/Wales/PHWC-47F369/2020|EPI_ISL_612665|2020 .................... .................... ..................... .................... .................... ....................
O 687 hCoV-19/Wales/PHWC-47F378/2020|EPI_ISL_612666|2020 .................... .................... ..................... .................... .................... ....................
O 688 hCoV-19/Wales/PHWC-47F387/2020|EPI_ISL_612667|2020 .................... .................... ..................... .................... .................... ....................
O 689 hCoV-19/Wales/PHWC-47F3B4/2020|EPI_ISL_612668|2020 .................... .................... ..................... .................... .................... ....................
O 690 hCoV-19/Wales/PHWC-47F3C3/2020|EPI_ISL_612669|2020 .................... .................... ..................... .................... .................... ....................
O 691 hCoV-19/Wales/PHWC-47F3D2/2020|EPI_ISL_612670|2020 .................... .................... ..................... .................... .................... ....................
O 692 hCoV-19/Wales/PHWC-47F3E1/2020|EPI_ISL_612671|2020 .................... .................... ..................... .................... .................... ....................
O 693 hCoV-19/Wales/PHWC-47F3F0/2020|EPI_ISL_612672|2020 .................... .................... ..................... .................... .................... ....................
O 694 hCoV-19/Wales/PHWC-47F40C/2020|EPI_ISL_612673|2020 .................... .................... ..................... .................... .................... ....................
O 695 hCoV-19/Wales/PHWC-47F41B/2020|EPI_ISL_612674|2020 .................... .................... ..................... .................... .................... ....................
O 696 hCoV-19/Wales/PHWC-47F439/2020|EPI_ISL_612675|2020 .................... .................... ..................... .................... .................... ....................
O 697 hCoV-19/Wales/PHWC-47F448/2020|EPI_ISL_612676|2020 .................... .................... ..................... .................... .................... ....................
O 698 hCoV-19/Wales/PHWC-47F457/2020|EPI_ISL_612677|2020 .................... .................... ..................... .................... .................... ....................
O 699 hCoV-19/Wales/PHWC-47F466/2020|EPI_ISL_612678|2020 .................... .................... ..................... .................... .................... ....................
O 700 hCoV-19/Wales/PHWC-47F475/2020|EPI_ISL_612679|2020 .................... .................... ..................... .................... .................... ....................
O 701 hCoV-19/Wales/PHWC-47F484/2020|EPI_ISL_612680|2020 .................... .................... ..................... .................... .................... ....................
O 702 hCoV-19/Wales/PHWC-47F493/2020|EPI_ISL_612681|2020 .................... .................... ..................... .................... .................... ....................
O 703 hCoV-19/Wales/PHWC-47F4A2/2020|EPI_ISL_612682|2020 .................... .................... ..................... .................... .................... ....................
O 704 hCoV-19/Wales/PHWC-47F4B1/2020|EPI_ISL_612683|2020 .................... .................... ..................... .................... .................... ....................
O 705 hCoV-19/Wales/PHWC-47F4C0/2020|EPI_ISL_612684|2020 .................... .................... ..................... .................... .................... ....................
O 706 hCoV-19/Wales/PHWC-47F4DF/2020|EPI_ISL_612685|2020 .................... .................... ..................... .................... .................... ....................
O 707 hCoV-19/Wales/PHWC-47F4EE/2020|EPI_ISL_612686|2020 .................... .................... ..................... .................... .................... ....................
O 708 hCoV-19/Wales/PHWC-47F509/2020|EPI_ISL_612687|2020 .................... .................... ..................... .................... .................... ....................
O 709 hCoV-19/Wales/PHWC-47F518/2020|EPI_ISL_612688|2020 .................... .................... ..................... .................... .................... ....................
O 710 hCoV-19/Wales/PHWC-47F527/2020|EPI_ISL_612689|2020 .................... .................... ..................... .................... .................... ....................
O 711 hCoV-19/Wales/PHWC-47F545/2020|EPI_ISL_612690|2020 .................... .................... ..................... .................... .................... ....................
O 712 hCoV-19/Wales/PHWC-47F554/2020|EPI_ISL_612691|2020 .................... .................... ..................... .................... .................... ....................
O 713 hCoV-19/Wales/PHWC-47F572/2020|EPI_ISL_612692|2020 .................... .................... ..................... .................... .................... ....................
O 714 hCoV-19/Wales/PHWC-47F581/2020|EPI_ISL_612693|2020 .................... .................... ..................... .................... .................... ....................
O 715 hCoV-19/Wales/PHWC-47F590/2020|EPI_ISL_612694|2020 .................... .................... ..................... .................... .................... ....................
O 716 hCoV-19/Wales/PHWC-47F5AF/2020|EPI_ISL_612695|2020 .................... .................... ..................... .................... .................... ....................
O 717 hCoV-19/Wales/PHWC-47F5BE/2020|EPI_ISL_612696|2020 .................... .................... ..................... .................... .................... ....................
O 718 hCoV-19/Wales/PHWC-47F5CD/2020|EPI_ISL_612697|2020 .................... .................... ..................... .................... .................... ....................
O 719 hCoV-19/Wales/PHWC-47F5DC/2020|EPI_ISL_612698|2020 .................... .................... ..................... .................... .................... ....................
O 720 hCoV-19/Wales/PHWC-47F5EB/2020|EPI_ISL_612699|2020 .................... .................... ..................... .................... .................... ....................
O 721 hCoV-19/USA/FL-BPHL-1803/2020|EPI_ISL_613900|2020- .................... .................... ..................... .................... .................... ....................
O 722 hCoV-19/USA/FL-BPHL-1804/2020|EPI_ISL_613901|2020- .................... .................... ..................... .................... .................... ....................
O 723 hCoV-19/USA/FL-BPHL-1806/2020|EPI_ISL_613902|2020- .................... .................... ..................... .................... .................... ....................
O 724 hCoV-19/USA/FL-BPHL-1807/2020|EPI_ISL_613903|2020- .................... .................... ..................... .................... .................... ....................
O 725 hCoV-19/USA/FL-BPHL-1815/2020|EPI_ISL_613904|2020- .................... .................... ..................... .................... .................... ....................
O 726 hCoV-19/USA/FL-BPHL-1816/2020|EPI_ISL_613905|2020- .................... .................... ..................... .................... .................... ....................
O 727 hCoV-19/USA/FL-BPHL-1817/2020|EPI_ISL_613906|2020- .................... .................... ..................... .................... .................... ....................
O 728 hCoV-19/USA/FL-BPHL-1818/2020|EPI_ISL_613907|2020- .................... .................... ..................... .................... .................... ....................
O 729 hCoV-19/USA/FL-BPHL-1820/2020|EPI_ISL_613908|2020- .................... .................... ..................... .................... .................... ....................
O 730 hCoV-19/USA/FL-BPHL-1822/2020|EPI_ISL_613909|2020- .................... .................... ..................... .................... .................... ....................
O 731 hCoV-19/USA/FL-BPHL-1823/2020|EPI_ISL_613910|2020- .................... .................... ..................... .................... .................... ....................
O 733 hCoV-19/USA/FL-BPHL-1825/2020|EPI_ISL_613912|2020- .................... .................... ..................... .................... .................... ....................
O 734 hCoV-19/USA/FL-BPHL-1827/2020|EPI_ISL_613913|2020- .................... .................... ..................... .................... .................... ....................
O 735 hCoV-19/USA/FL-BPHL-1828/2020|EPI_ISL_613914|2020- .................... .................... ..................... .................... .................... ....................
O 736 hCoV-19/USA/FL-BPHL-1830/2020|EPI_ISL_613915|2020- .................... .................... ..................... .................... .................... ....................
O 737 hCoV-19/USA/FL-BPHL-1833/2020|EPI_ISL_613916|2020- .................... .................... ..................... .................... .................... ....................
O 738 hCoV-19/USA/FL-BPHL-1834/2020|EPI_ISL_613917|2020- .................... .................... ..................... .................... .................... ....................
O 739 hCoV-19/USA/FL-BPHL-1837/2020|EPI_ISL_613918|2020- .................... .................... ..................... .................... .................... ....................
O 740 hCoV-19/USA/FL-BPHL-1839/2020|EPI_ISL_613919|2020- .................... .................... ..................... .................... .................... ....................
O 741 hCoV-19/USA/FL-BPHL-1840/2020|EPI_ISL_613920|2020- .................... .................... ..................... .................... .................... ....................
O 742 hCoV-19/USA/FL-BPHL-1841/2020|EPI_ISL_613921|2020- .................... .................... ..................... .................... .................... ....................
O 743 hCoV-19/USA/FL-BPHL-1842/2020|EPI_ISL_613922|2020- .................... .................... ..................... .................... .................... ....................
O 744 hCoV-19/USA/FL-BPHL-1843/2020|EPI_ISL_613923|2020- .................... .................... ..................... .................... .................... ....................
O 745 hCoV-19/USA/FL-BPHL-1844/2020|EPI_ISL_613924|2020- .................... .................... ..................... .................... .................... ....................
O 746 hCoV-19/USA/FL-BPHL-1845/2020|EPI_ISL_613925|2020- .................... .................... ..................... .................... .................... ....................
O 748 hCoV-19/USA/FL-BPHL-1847/2020|EPI_ISL_613927|2020- .................... .................... ..................... .................... .................... ....................
O 749 hCoV-19/USA/FL-BPHL-1848/2020|EPI_ISL_613928|2020- .................... .................... ..................... .................... .................... ....................
O 750 hCoV-19/USA/FL-BPHL-1849/2020|EPI_ISL_613929|2020- .................... .................... ..................... .................... .................... ....................
O 751 hCoV-19/USA/FL-BPHL-1850/2020|EPI_ISL_613930|2020- .................... .................... ..................... .................... .................... ....................
O 752 hCoV-19/USA/FL-BPHL-1851/2020|EPI_ISL_613931|2020- .................... .................... ..................... .................... .................... ....................
O 753 hCoV-19/USA/FL-BPHL-1852/2020|EPI_ISL_613932|2020- .................... .................... ..................... .................... .................... ....................
O 754 hCoV-19/USA/FL-BPHL-1853/2020|EPI_ISL_613933|2020- .................... .................... ..................... .................... .................... ....................
O 755 hCoV-19/USA/FL-BPHL-1854/2020|EPI_ISL_613934|2020- .................... .................... ..................... .................... .................... ....................
O 756 hCoV-19/USA/FL-BPHL-1855/2020|EPI_ISL_613935|2020- .................... .................... ..................... .................... .................... ....................
O 757 hCoV-19/USA/FL-BPHL-1858/2020|EPI_ISL_613936|2020- .................... .................... ..................... .................... .................... ....................
O 758 hCoV-19/USA/FL-BPHL-1859/2020|EPI_ISL_613937|2020- .................... .................... ..................... .................... .................... ....................
O 759 hCoV-19/USA/FL-BPHL-1860/2020|EPI_ISL_613938|2020- .................... .................... ..................... .................... .................... ....................

| | | | | | | | | |
|---|---|---|---|---|---|---|---|---|
| O | 760 | hCoV-19/USA/FL-BPHL-1861/2020\|EPI_ISL_613939\|2020- | ···················· | ···················· | ···················· | ···················· | ···················· | ···················· |
| O | 761 | hCoV-19/USA/FL-BPHL-1862/2020\|EPI_ISL_613940\|2020- | ···················· | ···················· | ···················· | ···················· | ···················· | ···················· |
| O | 762 | hCoV-19/USA/FL-BPHL-1863/2020\|EPI_ISL_613941\|2020- | ···················· | ···················· | ···················· | ···················· | ···················· | ···················· |
| O | 763 | hCoV-19/USA/FL-BPHL-1864/2020\|EPI_ISL_613942\|2020- | ···················· | ···················· | ···················· | ···················· | ···················· | ···················· |
| O | 764 | hCoV-19/USA/FL-BPHL-1865/2020\|EPI_ISL_613943\|2020- | ···················· | ···················· | ···················· | ···················· | ···················· | ···················· |
| O | 765 | hCoV-19/USA/FL-BPHL-1866/2020\|EPI_ISL_613944\|2020- | ···················· | ···················· | ···················· | ···················· | ···················· | ···················· |
| O | 766 | hCoV-19/USA/FL-BPHL-1931/2020\|EPI_ISL_613950\|2020- | ···················· | ···················· | ···················· | ···················· | ···················· | ···················· |
| O | 767 | hCoV-19/Brazil/SP-RP9/2020\|EPI_ISL_613951\|2020-04- | ···················· | ···················· | ···················· | ···················· | ···················· | ···················· |
| O | 768 | hCoV-19/Italy/SAR-ASSL-04/2020\|EPI_ISL_613955\|2020 | ···················· | ···················· | ···················· | ···················· | ···················· | ···················· |
| O | 769 | hCoV-19/England/MILK-A79A0A/2020\|EPI_ISL_606703\|20 | ···················· | ···················· | ···················· | ···················· | ···················· | ···················· |
| O | 770 | hCoV-19/England/MILK-A7A061/2020\|EPI_ISL_606704\|20 | ···················· | ···················· | ···················· | ···················· | ···················· | ···················· |
| O | 771 | hCoV-19/England/MILK-A7A3D1/2020\|EPI_ISL_606705\|20 | ···················· | ···················· | ···················· | ···················· | ···················· | ···················· |
| O | 772 | hCoV-19/England/MILK-A7ABCA/2020\|EPI_ISL_606706\|20 | ···················· | ···················· | ···················· | ···················· | ···················· | ···················· |
| O | 773 | hCoV-19/England/MILK-A7D040/2020\|EPI_ISL_606707\|20 | ···················· | ···················· | ···················· | ···················· | ···················· | ···················· |
| O | 774 | hCoV-19/England/MILK-A79BE9/2020\|EPI_ISL_606708\|20 | ···················· | ···················· | ···················· | ···················· | ···················· | ···················· |
| O | 775 | hCoV-19/England/MILK-A79B43/2020\|EPI_ISL_606709\|20 | ···················· | ···················· | ···················· | ···················· | ···················· | ···················· |
| O | 776 | hCoV-19/England/MILK-A79F56/2020\|EPI_ISL_606710\|20 | ···················· | ···················· | ···················· | ···················· | ···················· | ···················· |
| O | 777 | hCoV-19/England/MILK-A79D4D/2020\|EPI_ISL_606711\|20 | ···················· | ···················· | ···················· | ···················· | ···················· | ···················· |
| O | 778 | hCoV-19/England/MILK-A79B07/2020\|EPI_ISL_606712\|20 | ···················· | ···················· | ···················· | ···················· | ···················· | ···················· |
| O | 779 | hCoV-19/England/MILK-A79C40/2020\|EPI_ISL_606713\|20 | ···················· | ···················· | ···················· | ···················· | ···················· | ···················· |
| O | 780 | hCoV-19/England/MILK-A79BBC/2020\|EPI_ISL_606714\|20 | ···················· | ···················· | ···················· | ···················· | ···················· | ···················· |
| O | 781 | hCoV-19/England/MILK-A79879/2020\|EPI_ISL_606715\|20 | ···················· | ···················· | ···················· | ···················· | ···················· | ···················· |
| O | 782 | hCoV-19/England/MILK-A7A140/2020\|EPI_ISL_606716\|20 | ···················· | ···················· | ···················· | ···················· | ···················· | ···················· |
| O | 783 | hCoV-19/England/MILK-A798C4/2020\|EPI_ISL_606717\|20 | ···················· | ···················· | ···················· | ···················· | ······T············· | ···················· |
| O | 784 | hCoV-19/England/MILK-A79C13/2020\|EPI_ISL_606718\|20 | ···················· | ···················· | ···················· | ···················· | ······T············· | ···················· |
| O | 785 | hCoV-19/England/MILK-A7AB33/2020\|EPI_ISL_606719\|20 | ···················· | ···················· | ···················· | ···················· | ···················· | ···················· |
| O | 786 | hCoV-19/England/MILK-A7A1C8/2020\|EPI_ISL_606720\|20 | ···················· | ···················· | ···················· | ···················· | ······T············· | ···················· |
| O | 787 | hCoV-19/England/MILK-A7A131/2020\|EPI_ISL_606721\|20 | ···················· | ···················· | ···················· | ···················· | ···················· | ···················· |
| O | 788 | hCoV-19/England/MILK-A7A3B3/2020\|EPI_ISL_606722\|20 | ···················· | ···················· | ···················· | ···················· | ···················· | ···················· |
| O | 789 | hCoV-19/England/MILK-A7CE47/2020\|EPI_ISL_606723\|20 | ···················· | ···················· | ···················· | ···················· | ···················· | ···················· |
| O | 790 | hCoV-19/England/MILK-A7AC21/2020\|EPI_ISL_606724\|20 | ···················· | ···················· | ···················· | ···················· | ···················· | ···················· |
| O | 791 | hCoV-19/England/MILK-A7D0C8/2020\|EPI_ISL_606725\|20 | ···················· | ···················· | ···················· | ···················· | ······T············· | ···················· |
| O | 792 | hCoV-19/England/MILK-A7CFCC/2020\|EPI_ISL_606727\|20 | ···················· | ···················· | ···················· | ···················· | ···················· | ···················· |
| O | 793 | hCoV-19/England/MILK-A7A298/2020\|EPI_ISL_606729\|20 | ···················· | ···················· | ···················· | ···················· | ···················· | ···················· |
| O | 794 | hCoV-19/England/MILK-A7CE38/2020\|EPI_ISL_606731\|20 | ···················· | ···················· | ···················· | ···················· | ···················· | ···················· |
| O | 795 | hCoV-19/England/MILK-A7A5DB/2020\|EPI_ISL_606734\|20 | ···················· | ···················· | ···················· | ···················· | ······T············· | ···················· |
| O | 796 | hCoV-19/England/MILK-A7A27A/2020\|EPI_ISL_606735\|20 | ···················· | ···················· | ···················· | ···················· | ···················· | ···················· |
| O | 797 | hCoV-19/England/MILK-A79CD7/2020\|EPI_ISL_606736\|20 | ···················· | ···················· | ···················· | ···················· | ···················· | ···················· |
| O | 798 | hCoV-19/England/MILK-A796CA/2020\|EPI_ISL_606737\|20 | ···················· | ·A·················· | ···················· | ···················· | ···················· | ···A················ |
| O | 799 | hCoV-19/England/MILK-A79712/2020\|EPI_ISL_606738\|20 | ···················· | ···················· | ···················· | ···················· | ···················· | ···················· |
| O | 800 | hCoV-19/England/MILK-A7A034/2020\|EPI_ISL_606739\|20 | ···················· | ···················· | ···················· | ···················· | ···················· | ···················· |
| O | 801 | hCoV-19/England/MILK-A7ACA9/2020\|EPI_ISL_606741\|20 | ···················· | ···················· | ···················· | ···················· | ···················· | ···················· |
| O | 802 | hCoV-19/England/MILK-A7D031/2020\|EPI_ISL_606743\|20 | ···················· | ···················· | ···················· | ···················· | ···················· | ···················· |
| O | 803 | hCoV-19/England/MILK-A7A8F0/2020\|EPI_ISL_606744\|20 | ···················· | ···················· | ···················· | ···················· | ···················· | ···················· |
| O | 804 | hCoV-19/England/MILK-A7A368/2020\|EPI_ISL_606746\|20 | ···················· | ···················· | ···················· | ···················· | ···················· | ···················· |
| O | 805 | hCoV-19/England/MILK-A79BF8/2020\|EPI_ISL_606748\|20 | ···················· | ···················· | ···················· | ···················· | ···················· | ···················· |
| O | 806 | hCoV-19/England/MILK-A7984C/2020\|EPI_ISL_606750\|20 | ···················· | ···················· | ···················· | ···················· | ···················· | ···················· |
| O | 807 | hCoV-19/England/MILK-A7CF17/2020\|EPI_ISL_606751\|20 | ···················· | ···················· | ···················· | ···················· | ···················· | ···················· |
| O | 808 | hCoV-19/England/MILK-A7A1B9/2020\|EPI_ISL_606752\|20 | ···················· | ···················· | ···················· | ···················· | ···················· | ···················· |
| O | 809 | hCoV-19/England/MILK-A79A55/2020\|EPI_ISL_606753\|20 | ···················· | ···················· | ···················· | ···················· | ···················· | ···················· |
| O | 810 | hCoV-19/England/MILK-A7A5CC/2020\|EPI_ISL_606755\|20 | ···················· | ···················· | ···················· | ···················· | ···················· | ···················· |
| O | 811 | hCoV-19/England/MILK-A79E59/2020\|EPI_ISL_606756\|20 | ···················· | ···················· | ···················· | ···················· | ···················· | ···················· |
| O | 812 | hCoV-19/England/MILK-A7A2C5/2020\|EPI_ISL_606757\|20 | ···················· | ···················· | ···················· | ···················· | ···················· | ···················· |
| O | 813 | hCoV-19/England/MILK-A7CE74/2020\|EPI_ISL_606758\|20 | ···················· | ···················· | ···················· | ···················· | ···················· | ···················· |
| O | 814 | hCoV-19/England/MILK-A79888/2020\|EPI_ISL_606759\|20 | ···················· | ·A·················· | ···················· | ···················· | ···················· | ···A················ |
| O | 815 | hCoV-19/England/MILK-A79FED/2020\|EPI_ISL_606760\|20 | ···················· | ···················· | ···················· | ···················· | ···················· | ···················· |
| O | 816 | hCoV-19/England/MILK-A7CF71/2020\|EPI_ISL_606761\|20 | ···················· | ···················· | ···················· | ···················· | ······T············· | ···················· |
| O | 817 | hCoV-19/England/MILK-A79624/2020\|EPI_ISL_606762\|20 | ···················· | ···················· | ···················· | ···················· | ···················· | ···················· |
| O | 818 | hCoV-19/England/MILK-A79EFF/2020\|EPI_ISL_606763\|20 | ···················· | ···················· | ···················· | ···················· | ···················· | ···················· |
| O | 819 | hCoV-19/England/MILK-A79CAA/2020\|EPI_ISL_606765\|20 | ···················· | ···················· | ···················· | ···················· | ···················· | ···················· |
| O | 820 | hCoV-19/England/MILK-A79FDE/2020\|EPI_ISL_606766\|20 | ···················· | ···················· | ···················· | ···················· | ···················· | ···················· |
| O | 821 | hCoV-19/England/MILK-A7CD3B/2020\|EPI_ISL_606767\|20 | ···················· | ···················· | ···················· | ···················· | ···················· | ···················· |
| O | 822 | hCoV-19/England/MILK-A795DC/2020\|EPI_ISL_606768\|20 | ···················· | ···················· | ···················· | ···················· | ···················· | ···················· |
| O | 823 | hCoV-19/England/MILK-A7CEFC/2020\|EPI_ISL_606769\|20 | ···················· | ···················· | ···················· | ···················· | ···················· | ···················· |
| O | 824 | hCoV-19/England/MILK-A7A8D2/2020\|EPI_ISL_606770\|20 | ···················· | ·A·················· | ···················· | ···················· | ···················· | ···A················ |
| O | 825 | hCoV-19/England/MILK-A7A4DE/2020\|EPI_ISL_606771\|20 | ···················· | ···················· | ···················· | ···················· | ···················· | ···················· |
| O | 826 | hCoV-19/England/MILK-A79EE0/2020\|EPI_ISL_606772\|20 | ···················· | ···················· | ···················· | ···················· | ······T············· | ···················· |
| O | 827 | hCoV-19/England/MILK-A7A429/2020\|EPI_ISL_606773\|20 | ···················· | ···················· | ···················· | ···················· | ···················· | ···················· |
| O | 828 | hCoV-19/England/MILK-A7A0CB/2020\|EPI_ISL_606775\|20 | ···················· | ···················· | ···················· | ···················· | ···················· | ···················· |
| O | 829 | hCoV-19/England/MILK-A7979A/2020\|EPI_ISL_606776\|20 | ···················· | ···················· | ···················· | ···················· | ···················· | ···················· |
| O | 830 | hCoV-19/England/MILK-A796E8/2020\|EPI_ISL_606777\|20 | ···················· | ·A·················· | ···················· | ···················· | ···················· | ···················· |
| O | 832 | hCoV-19/England/MILK-A797C7/2020\|EPI_ISL_606779\|20 | ···················· | ·A·················· | ···················· | ···················· | ···················· | ···················· |
| O | 833 | hCoV-19/England/MILK-A795FA/2020\|EPI_ISL_606780\|20 | ···················· | ···················· | ···················· | ···················· | ···················· | ···················· |
| O | 834 | hCoV-19/England/MILK-A79F74/2020\|EPI_ISL_606781\|20 | ···················· | ·A·······A·········· | ···················· | ···················· | ···················· | ···················· |
| O | 835 | hCoV-19/England/MILK-A79D01/2020\|EPI_ISL_606782\|20 | ···················· | ···················· | ···················· | ···················· | ···················· | ···················· |
| O | 836 | hCoV-19/England/MILK-A7992B/2020\|EPI_ISL_606783\|20 | ···················· | ···················· | ···················· | ···················· | ···················· | ···················· |
| O | 837 | hCoV-19/England/MILK-A7A23E/2020\|EPI_ISL_606784\|20 | ···················· | ···················· | ···················· | ···················· | ···················· | ···················· |
| O | 838 | hCoV-19/England/MILK-A79F47/2020\|EPI_ISL_606785\|20 | ···················· | ···················· | ···················· | ···················· | ···················· | ···················· |
| O | 839 | hCoV-19/England/MILK-A79CE6/2020\|EPI_ISL_606787\|20 | ···················· | ···················· | ···················· | ···················· | ···················· | ···················· |
| O | 840 | hCoV-19/England/MILK-A79EC2/2020\|EPI_ISL_606788\|20 | ···················· | ···················· | ···················· | ···················· | ···················· | ···················· |

| | | | | | | | |
|---|---|---|---|---|---|---|---|
| O | 842 hCoV-19/England/MILK-A79C31/2020\|EPI_ISL_606790\|20 | ···················· | ···················· | ···················· | ···················· | ···················· | ···················· |
| O | 843 hCoV-19/England/MILK-A7CE92/2020\|EPI_ISL_606792\|20 | ···················· | ···················· | ···················· | ···················· | ······T············· | ···················· |
| O | 844 hCoV-19/England/MILK-A7D05F/2020\|EPI_ISL_606793\|20 | ···················· | ·A·················· | ···················· | ···················· | ···················· | ···A················ |
| O | 845 hCoV-19/England/MILK-A79C8C/2020\|EPI_ISL_606796\|20 | ···················· | ···················· | ···················· | ···················· | ···················· | ···················· |
| O | 846 hCoV-19/England/MILK-A7A32C/2020\|EPI_ISL_606798\|20 | ···················· | ·A·················· | ···················· | ···················· | ···················· | ···················· |
| O | 847 hCoV-19/England/MILK-A797D6/2020\|EPI_ISL_606799\|20 | ···················· | ···················· | ···················· | ···················· | ···················· | ···················· |
| O | 848 hCoV-19/England/ALDP-A695DF/2020\|EPI_ISL_609000\|20 | ···················· | ···················· | ···················· | ···················· | ···················· | ···················· |
| O | 849 hCoV-19/England/MILK-A6BCE7/2020\|EPI_ISL_609001\|20 | ···················· | ···················· | ···················· | ···················· | ···················· | ···················· |
| O | 850 hCoV-19/England/ALDP-A67FD3/2020\|EPI_ISL_609002\|20 | ···················· | ···················· | ···················· | ···················· | ···················· | ···················· |
| O | 851 hCoV-19/England/CAMC-A64C84/2020\|EPI_ISL_609003\|20 | ···················· | ·A·················· | ···················· | ···················· | ···················· | ···A················ |
| O | 852 hCoV-19/England/ALDP-A6937B/2020\|EPI_ISL_609004\|20 | ···················· | ···················· | ···················· | ···················· | ···················· | ···················· |
| O | 853 hCoV-19/England/MILK-A6C28A/2020\|EPI_ISL_609006\|20 | ···················· | ···················· | ···················· | ···················· | ···················· | ···················· |
| O | 854 hCoV-19/England/ALDP-A69627/2020\|EPI_ISL_609007\|20 | ···················· | ···················· | ···················· | ···················· | ···················· | ···················· |
| O | 855 hCoV-19/England/MILK-A6C105/2020\|EPI_ISL_609008\|20 | ···················· | ···················· | ···················· | ···················· | ···················· | ···················· |
| O | 856 hCoV-19/England/ALDP-A6827F/2020\|EPI_ISL_609009\|20 | ···················· | ···················· | ···················· | ···················· | ···················· | ···················· |
| O | 857 hCoV-19/England/ALDP-A93D89/2020\|EPI_ISL_611206\|20 | ···················· | ·A·················· | ···················· | ···················· | ···················· | ···A················ |
| O | 858 hCoV-19/England/ALDP-A93606/2020\|EPI_ISL_611207\|20 | ···················· | ·A·················· | ···················· | ···················· | ···················· | ···A················ |
| O | 859 hCoV-19/England/ALDP-A93F1A/2020\|EPI_ISL_611208\|20 | ···················· | ···················· | ···················· | ···················· | ···················· | ···················· |
| O | 860 hCoV-19/England/ALDP-A9290E/2020\|EPI_ISL_611209\|20 | ···················· | ···················· | ···················· | ···················· | ···················· | ···················· |
| O | 861 hCoV-19/England/ALDP-A94395/2020\|EPI_ISL_611210\|20 | ···················· | ···················· | ···················· | ···················· | ···················· | ···················· |
| O | 862 hCoV-19/England/ALDP-A948A5/2020\|EPI_ISL_611211\|20 | ···················· | ···················· | ···················· | ···················· | ···················· | ···················· |
| O | 863 hCoV-19/England/ALDP-A936CA/2020\|EPI_ISL_611212\|20 | ···················· | ···················· | ···················· | ···················· | ···················· | ···················· |
| O | 864 hCoV-19/England/ALDP-A92DE4/2020\|EPI_ISL_611213\|20 | ···················· | ·A·················· | ···················· | ···················· | ···················· | ···················· |
| O | 865 hCoV-19/England/ALDP-A9283E/2020\|EPI_ISL_611214\|20 | ···················· | ···················· | ···················· | ···················· | ···················· | ···················· |
| O | 866 hCoV-19/England/ALDP-A9481E/2020\|EPI_ISL_611216\|20 | ···················· | ···················· | ···················· | ···················· | ···················· | ···················· |
| O | 867 hCoV-19/England/ALDP-A9250A/2020\|EPI_ISL_611220\|20 | ···················· | ···················· | ···················· | ···················· | ···················· | ···················· |
| O | 868 hCoV-19/England/ALDP-A9232E/2020\|EPI_ISL_611221\|20 | ···················· | ···················· | ···················· | ···················· | ···················· | ···················· |
| O | 869 hCoV-19/England/ALDP-A93DC5/2020\|EPI_ISL_611250\|20 | ···················· | ···················· | ···················· | ···················· | ···················· | ···················· |
| O | 870 hCoV-19/England/ALDP-A929B3/2020\|EPI_ISL_611251\|20 | ···················· | ···················· | ···················· | ···················· | ···················· | ···················· |
| O | 871 hCoV-19/England/ALDP-A925FB/2020\|EPI_ISL_611252\|20 | ···················· | ···················· | ···················· | ···················· | ···················· | ···················· |
| O | 872 hCoV-19/England/ALDP-A924EF/2020\|EPI_ISL_611253\|20 | ···················· | ···················· | ···················· | ···················· | ···················· | ···················· |
| O | 873 hCoV-19/England/ALDP-A939D0/2020\|EPI_ISL_611255\|20 | ···················· | ···················· | ···················· | ···················· | ···················· | ···················· |
| O | 874 hCoV-19/England/ALDP-A93EB3/2020\|EPI_ISL_611256\|20 | ···················· | ···················· | ···················· | ···················· | ···················· | ···················· |
| O | 875 hCoV-19/England/ALDP-A92A29/2020\|EPI_ISL_611258\|20 | ···················· | ···················· | ···················· | ···················· | ···················· | ···················· |
| O | 876 hCoV-19/England/ALDP-A926E9/2020\|EPI_ISL_611259\|20 | ···················· | ···················· | ···················· | ···················· | ···················· | ···················· |
| O | 877 hCoV-19/England/ALDP-A943C2/2020\|EPI_ISL_611260\|20 | ···················· | ···················· | ···················· | ···················· | ···················· | ···················· |
| O | 878 hCoV-19/England/ALDP-A93BCB/2020\|EPI_ISL_611261\|20 | ···················· | ···················· | ···················· | ···················· | ···················· | ···················· |
| O | 879 hCoV-19/England/ALDP-A9441A/2020\|EPI_ISL_611262\|20 | ···················· | ·A·················· | ···················· | ···················· | ···················· | ···A················ |
| O | 880 hCoV-19/England/ALDP-A93E0E/2020\|EPI_ISL_611263\|20 | ···················· | ···················· | ···················· | ···················· | ···················· | ···················· |
| O | 881 hCoV-19/England/ALDP-A92DB7/2020\|EPI_ISL_611264\|20 | ···················· | ···················· | ···················· | ···················· | ···················· | ···················· |
| O | 882 hCoV-19/England/ALDP-A92986/2020\|EPI_ISL_611265\|20 | ···················· | ···················· | ···················· | ···················· | ···················· | ···················· |
| O | 883 hCoV-19/England/ALDP-A94052/2020\|EPI_ISL_611266\|20 | ···················· | ···················· | ···················· | ···················· | ···················· | ···················· |
| O | 884 hCoV-19/England/ALDP-A939B2/2020\|EPI_ISL_611267\|20 | ···················· | ···················· | ···················· | ···················· | ···················· | ···················· |
| O | 885 hCoV-19/England/ALDP-A92D99/2020\|EPI_ISL_611268\|20 | ···················· | ···················· | ···················· | ···················· | ···················· | ···················· |
| O | 886 hCoV-19/England/ALDP-A93AFB/2020\|EPI_ISL_611269\|20 | ···················· | ···················· | ···················· | ···················· | ···················· | ···················· |
| O | 887 hCoV-19/England/ALDP-A93D2F/2020\|EPI_ISL_611270\|20 | ···················· | ···················· | ···················· | ···················· | ···················· | ···················· |
| O | 888 hCoV-19/England/ALDP-A92C9C/2020\|EPI_ISL_611271\|20 | ···················· | ···················· | ···················· | ···················· | ···················· | ···················· |
| O | 889 hCoV-19/England/ALDP-A9233D/2020\|EPI_ISL_611272\|20 | ···················· | ···················· | ···················· | ···················· | ···················· | ···················· |
| O | 890 hCoV-19/England/ALDP-A928A7/2020\|EPI_ISL_611273\|20 | ···················· | ···················· | ···················· | ···················· | ···················· | ···················· |
| O | 891 hCoV-19/England/ALDP-A92BBD/2020\|EPI_ISL_611275\|20 | ···················· | ···················· | ···················· | ···················· | ···················· | ···················· |
| O | 892 hCoV-19/England/ALDP-A93897/2020\|EPI_ISL_611277\|20 | ···················· | ···················· | ···················· | ···················· | ···················· | ···················· |
| O | 893 hCoV-19/England/ALDP-A92625/2020\|EPI_ISL_611278\|20 | ···················· | ···················· | ···················· | ···················· | ···················· | ···················· |
| O | 894 hCoV-19/England/ALDP-A93B25/2020\|EPI_ISL_611279\|20 | ···················· | ···················· | ···················· | ···················· | ···················· | ···················· |
| O | 895 hCoV-19/England/ALDP-A93EE0/2020\|EPI_ISL_611280\|20 | ···················· | ·A·················· | ···················· | ···················· | ···················· | ···················· |
| O | 896 hCoV-19/England/ALDP-A92B80/2020\|EPI_ISL_611281\|20 | ···················· | ···················· | ···················· | ···················· | ···················· | ···················· |
| O | 897 hCoV-19/England/ALDP-A923A6/2020\|EPI_ISL_611282\|20 | ···················· | ···················· | ···················· | ···················· | ···················· | ···················· |
| O | 898 hCoV-19/England/ALDP-A943A4/2020\|EPI_ISL_611283\|20 | ···················· | ···················· | ···················· | ···················· | ···················· | ···················· |
| O | 899 hCoV-19/England/ALDP-A92D8A/2020\|EPI_ISL_611284\|20 | ···················· | ···················· | ···················· | ···················· | ···················· | ···················· |
| O | 900 hCoV-19/England/ALDP-A935DC/2020\|EPI_ISL_611285\|20 | ···················· | ···················· | ···················· | ···················· | ···················· | ···················· |
| O | 901 hCoV-19/England/ALDP-A92EA5/2020\|EPI_ISL_611286\|20 | ···················· | ···················· | ···················· | ···················· | ···················· | ···················· |
| O | 902 hCoV-19/England/QEUH-A9F594/2020\|EPI_ISL_611287\|20 | ···················· | ···················· | ···················· | ···················· | ···················· | ···················· |
| O | 903 hCoV-19/England/QEUH-A9F637/2020\|EPI_ISL_611288\|20 | ···················· | ···················· | ···················· | ···················· | ···················· | ···················· |
| O | 904 hCoV-19/England/QEUH-A9EFB5/2020\|EPI_ISL_611289\|20 | ···················· | ·A·················· | ···················· | ···················· | ···················· | ···················· |
| O | 905 hCoV-19/England/QEUH-A9FB0B/2020\|EPI_ISL_611290\|20 | ···················· | ···················· | ···················· | ···················· | ···················· | ···················· |
| O | 906 hCoV-19/England/QEUH-A9F3B8/2020\|EPI_ISL_611291\|20 | ···················· | ···················· | ···················· | ···················· | ···················· | ···················· |
| O | 907 hCoV-19/England/QEUH-A9B677/2020\|EPI_ISL_611292\|20 | ···················· | ···················· | ···················· | ···················· | ···················· | ···················· |
| O | 908 hCoV-19/England/QEUH-A9E9E4/2020\|EPI_ISL_611293\|20 | ···················· | ···················· | ···················· | ···················· | ···················· | ···················· |
| O | 909 hCoV-19/England/QEUH-AA0299/2020\|EPI_ISL_611294\|20 | ···················· | ···················· | ···················· | ···················· | ···················· | ···················· |
| O | 910 hCoV-19/England/QEUH-A9ADCE/2020\|EPI_ISL_611295\|20 | ···················· | ·A·················· | ···················· | ···················· | ···················· | ···················· |
| O | 911 hCoV-19/England/QEUH-A9EC81/2020\|EPI_ISL_611296\|20 | ···················· | ···················· | ···················· | ···················· | ···················· | ···················· |
| O | 912 hCoV-19/England/QEUH-A9F6EC/2020\|EPI_ISL_611297\|20 | ···················· | ···················· | ···················· | ···················· | ···················· | ···················· |
| O | 913 hCoV-19/England/QEUH-A9ED60/2020\|EPI_ISL_611299\|20 | ···················· | ···················· | ···················· | ···················· | ···················· | ···················· |
| O | 914 hCoV-19/England/QEUH-A9F51C/2020\|EPI_ISL_611300\|20 | ···················· | ···················· | ···················· | ···················· | ···················· | ···················· |
| O | 915 hCoV-19/England/QEUH-A9FE6C/2020\|EPI_ISL_611301\|20 | ···················· | ···················· | ···················· | ···················· | ···················· | ···················· |
| O | 916 hCoV-19/England/QEUH-A9AE16/2020\|EPI_ISL_611302\|20 | ···················· | ···················· | ···················· | ···················· | ···················· | ···················· |
| O | 917 hCoV-19/England/QEUH-A9FB65/2020\|EPI_ISL_611305\|20 | ···················· | ···················· | ···················· | ···················· | ···················· | ···················· |
| O | 918 hCoV-19/England/QEUH-A9B6B3/2020\|EPI_ISL_611306\|20 | ···················· | ···················· | ···················· | ···················· | ···················· | ···················· |
| O | 919 hCoV-19/England/QEUH-A9EE21/2020\|EPI_ISL_611307\|20 | ···················· | ···················· | ···················· | ···················· | ···················· | ···················· |
| O | 920 hCoV-19/England/QEUH-A9F682/2020\|EPI_ISL_611308\|20 | ···················· | ···················· | ···················· | ···················· | ···················· | ···················· |
| O | 921 hCoV-19/England/QEUH-A9FB83/2020\|EPI_ISL_611309\|20 | ···················· | ···················· | ···················· | ···················· | ······T············· | ···················· |

O 922 hCoV-19/England/QEUH-A9F646/2020|EPI_ISL_611310|20 ···················· ···················· ···················· ···················· ···················· ····················
O 923 hCoV-19/England/QEUH-AA0062/2020|EPI_ISL_611312|20 ···················· ···················· ···················· ···················· ···················· ····················
O 924 hCoV-19/England/QEUH-AA02D5/2020|EPI_ISL_611314|20 ···················· ···················· ···················· ···················· ···················· ····················
O 925 hCoV-19/England/QEUH-A9EBB1/2020|EPI_ISL_611315|20 ···················· ·A·················· ···················· ···················· ···················· ··A·················
O 926 hCoV-19/England/QEUH-A9ED9D/2020|EPI_ISL_611317|20 ···················· ···················· ···················· ···················· ···················· ····················
O 927 hCoV-19/England/QEUH-A9B756/2020|EPI_ISL_611320|20 ···················· ···················· ···················· ···················· ···················· ····················
O 928 hCoV-19/England/QEUH-AA0202/2020|EPI_ISL_611321|20 ···················· ···················· ···················· ···················· ···················· ····················
O 929 hCoV-19/England/QEUH-A9E911/2020|EPI_ISL_611322|20 ···················· ···················· ···················· ···················· ···················· ····················
O 930 hCoV-19/England/QEUH-A9EF1F/2020|EPI_ISL_611323|20 ···················· ···················· ···················· ···················· ···················· ····················
O 931 hCoV-19/England/QEUH-A9EFA6/2020|EPI_ISL_611324|20 ···················· ···················· ···················· ···················· ···················· ····················
O 932 hCoV-19/England/QEUH-A9EDD9/2020|EPI_ISL_611325|20 ···················· ···················· ···················· ···················· ···················· ····················
O 933 hCoV-19/England/MILK-A22A00/2020|EPI_ISL_612537|20 ···················· ···················· ···················· ···················· ···················· ····················
O 934 hCoV-19/England/MILK-A22A2E/2020|EPI_ISL_612538|20 ···················· ···················· ···················· ···················· ···················· ····················
O 935 hCoV-19/England/MILK-A22A4C/2020|EPI_ISL_612539|20 ···················· ···················· ···················· ···················· ···················· ····················
O 936 hCoV-19/England/MILK-A22A79/2020|EPI_ISL_612540|20 ···················· ···················· ···················· ···················· ···················· ····················
O 937 hCoV-19/England/MILK-A22A88/2020|EPI_ISL_612541|20 ···················· ···················· ···················· ···················· ···················· ····················
O 938 hCoV-19/England/MILK-A22AA6/2020|EPI_ISL_612542|20 ···················· ···················· ···················· ···················· ···················· ····················
O 939 hCoV-19/England/MILK-A22AB5/2020|EPI_ISL_612543|20 ···················· ···················· ···················· ···················· ······T············· ····················
O 940 hCoV-19/England/MILK-A22AD3/2020|EPI_ISL_612544|20 ···················· ···················· ···················· ···················· ···················· ····················
O 941 hCoV-19/England/MILK-A22AF1/2020|EPI_ISL_612545|20 ···················· ···················· ···················· ···················· ···················· ····················
O 943 hCoV-19/Northern Ireland/NIRE-108EC1/2020|EPI_ISL_ ···················· ···················· ···················· ···················· ···················· ····················
O 944 hCoV-19/England/NORT-1B6BF17/2020|EPI_ISL_612553|2 ···················· ···················· ···················· ···················· ···················· ····················
O 945 hCoV-19/England/NOTT-1176D6/2020|EPI_ISL_612580|20 ···················· ···················· ···················· ···················· ···················· ····················
O 946 hCoV-19/England/NOTT-1176F4/2020|EPI_ISL_612581|20 ···················· ···················· ···················· ···················· ···················· ····················
O 947 hCoV-19/England/NOTT-117700/2020|EPI_ISL_612582|20 ···················· ···················· ···················· ···················· ···················· ····················
O 948 hCoV-19/England/NOTT-11771F/2020|EPI_ISL_612583|20 ···················· ···················· ···················· ···················· ···················· ····················
O 949 hCoV-19/England/NOTT-11772E/2020|EPI_ISL_612584|20 ···················· ···················· ···················· ···················· ···················· ····················
O 950 hCoV-19/England/NOTT-11776A/2020|EPI_ISL_612585|20 ···················· ···················· ···················· ···················· ···················· ····················
O 952 hCoV-19/England/NOTT-117C3E/2020|EPI_ISL_612587|20 ···················· ···················· ···················· ···················· ···················· ····················
O 953 hCoV-19/England/NOTT-117C4D/2020|EPI_ISL_612588|20 ···················· ···················· ···················· ···················· ···················· ····················
O 954 hCoV-19/England/NOTT-117C6B/2020|EPI_ISL_612589|20 ···················· ···················· ···················· ···················· ···················· ····················
O 955 hCoV-19/England/NOTT-117D95/2020|EPI_ISL_612590|20 ···················· ···················· ···················· ···················· ···················· ····················
O 956 hCoV-19/England/NOTT-117F44/2020|EPI_ISL_612591|20 ···················· ···················· ···················· ···················· ···················· ····················
O 957 hCoV-19/England/NOTT-117F80/2020|EPI_ISL_612592|20 ···················· ···················· ···················· ···················· ···················· ····················
O 958 hCoV-19/England/NOTT-117F9F/2020|EPI_ISL_612593|20 ···················· ···················· ···················· ···················· ···················· ····················
O 959 hCoV-19/England/NOTT-117FAE/2020|EPI_ISL_612594|20 ···················· ···················· ···················· ···················· ···················· ····················
O 960 hCoV-19/England/NOTT-11816B/2020|EPI_ISL_612595|20 ···················· ···················· ···················· ···················· ···················· ····················
O 961 hCoV-19/England/NOTT-11817A/2020|EPI_ISL_612596|20 ···················· ···················· ···················· ···················· ···················· ····················
O 962 hCoV-19/England/NOTT-118189/2020|EPI_ISL_612597|20 ···················· ···················· ···················· ···················· ···················· ····················
O 963 hCoV-19/England/NOTT-1181A7/2020|EPI_ISL_612598|20 ···················· ···················· ···················· ···················· ···················· ····················
O 964 hCoV-19/England/NOTT-1183B0/2020|EPI_ISL_612599|20 ···················· ···················· ···················· ···················· ···················· ····················
O 966 hCoV-19/Wales/PHWC-47EFEE/2020|EPI_ISL_612615|2020 ···················· ···················· ···················· ···················· ···················· ····················
O 967 hCoV-19/Wales/PHWC-47F008/2020|EPI_ISL_612616|2020 ···················· ···················· ···················· ···················· ···················· ····················
O 968 hCoV-19/Wales/PHWC-47F017/2020|EPI_ISL_612617|2020 ···················· ···················· ···················· ···················· ···················· ····················
O 969 hCoV-19/Wales/PHWC-47F026/2020|EPI_ISL_612618|2020 ···················· ···················· ···················· ···················· ···················· ····················
O 970 hCoV-19/Wales/PHWC-47F035/2020|EPI_ISL_612619|2020 ···················· ···················· ···················· ···················· ···················· ····················
O 971 hCoV-19/Wales/PHWC-47F062/2020|EPI_ISL_612622|2020 ···················· ···················· ···················· ···················· ···················· ····················
O 972 hCoV-19/Wales/PHWC-47F09F/2020|EPI_ISL_612625|2020 ···················· ···················· ···················· ···················· ···················· ····················
O 974 hCoV-19/Wales/PHWC-47F0DB/2020|EPI_ISL_612627|2020 ···················· ···················· ···················· ···················· ···················· ····················
O 975 hCoV-19/Wales/PHWC-47F0EA/2020|EPI_ISL_612628|2020 ···················· ···················· ···················· ···················· ···················· ····················
O 976 hCoV-19/Wales/PHWC-47F0F9/2020|EPI_ISL_612629|2020 ···················· ···················· ···················· ···················· ···················· ····················
O 977 hCoV-19/Wales/PHWC-47F105/2020|EPI_ISL_612630|2020 ···················· ···················· ···················· ···················· ···················· ····················
O 978 hCoV-19/Wales/PHWC-47F114/2020|EPI_ISL_612631|2020 ···················· ···················· ···················· ···················· ···················· ····················
O 979 hCoV-19/Wales/PHWC-47F123/2020|EPI_ISL_612632|2020 ···················· ···················· ···················· ···················· ···················· ····················
O 980 hCoV-19/Wales/PHWC-47F132/2020|EPI_ISL_612633|2020 ···················· ···················· ···················· ···················· ···················· ····················
O 981 hCoV-19/Wales/PHWC-47F141/2020|EPI_ISL_612634|2020 ···················· ···················· ···················· ···················· ···················· ····················
O 982 hCoV-19/Wales/PHWC-47F150/2020|EPI_ISL_612635|2020 ···················· ···················· ···················· ···················· ···················· ····················
O 983 hCoV-19/Wales/PHWC-47F16F/2020|EPI_ISL_612636|2020 ···················· ···················· ···················· ···················· ···················· ····················
O 984 hCoV-19/Wales/PHWC-47F17E/2020|EPI_ISL_612637|2020 ···················· ···················· ···················· ···················· ···················· ····················
O 985 hCoV-19/Wales/PHWC-47F18D/2020|EPI_ISL_612638|2020 ···················· ···················· ···················· ···················· ···················· ····················
O 986 hCoV-19/Wales/PHWC-47F19C/2020|EPI_ISL_612639|2020 ···················· ···················· ···················· ···················· ···················· ····················
O 987 hCoV-19/Wales/PHWC-47F1AB/2020|EPI_ISL_612640|2020 ···················· ···················· ···················· ···················· ···················· ····················
O 988 hCoV-19/Wales/PHWC-47F1BA/2020|EPI_ISL_612641|2020 ···················· ···················· ···················· ···················· ···················· ····················
O 989 hCoV-19/Wales/PHWC-47F1C9/2020|EPI_ISL_612642|2020 ···················· ···················· ···················· ···················· ···················· ····················
O 990 hCoV-19/Wales/PHWC-47F1D8/2020|EPI_ISL_612643|2020 ···················· ···················· ···················· ···················· ···················· ····················
O 991 hCoV-19/Wales/PHWC-47F1F6/2020|EPI_ISL_612644|2020 ···················· ···················· ···················· ···················· ···················· ····················
O 992 hCoV-19/Wales/PHWC-47F202/2020|EPI_ISL_612645|2020 ···················· ···················· ···················· ···················· ···················· ····················
O 993 hCoV-19/Wales/PHWC-47F211/2020|EPI_ISL_612646|2020 ···················· ···················· ···················· ···················· ···················· ····················
O 994 hCoV-19/Wales/PHWC-47F220/2020|EPI_ISL_612647|2020 ···················· ···················· ···················· ···················· ···················· ····················
O 995 hCoV-19/Wales/PHWC-47F23F/2020|EPI_ISL_612648|2020 ···················· ···················· ···················· ···················· ···················· ····················
O 996 hCoV-19/Wales/PHWC-47F24E/2020|EPI_ISL_612649|2020 ···················· ···················· ···················· ···················· ···················· ····················
O 997 hCoV-19/Wales/PHWC-47F25D/2020|EPI_ISL_612650|2020 ···················· ···················· ···················· ···················· ···················· ····················
O 998 hCoV-19/Wales/PHWC-47F26C/2020|EPI_ISL_612651|2020 ···················· ···················· ···················· ···················· ···················· ····················
O 999 hCoV-19/Wales/PHWC-47F27B/2020|EPI_ISL_612652|2020 ···················· ···················· ···················· ···················· ···················· ····················
O 1000 hCoV-19/Wales/PHWC-47F28A/2020|EPI_ISL_612653|2020 ···················· ···················· ···················· ···················· ···················· ····················
O 1001 hCoV-19/Wales/PHWC-47F299/2020|EPI_ISL_612654|2020 ···················· ···················· ···················· ···················· ···················· ····················
O 1002 hCoV-19/Wales/PHWC-47F2A8/2020|EPI_ISL_612655|2020 ···················· ···················· ···················· ···················· ···················· ····················
O 1003 hCoV-19/Wales/PHWC-47F2B7/2020|EPI_ISL_612656|2020 ···················· ···················· ···················· ···················· ···················· ····················
O 1004 hCoV-19/Wales/PHWC-47F2C6/2020|EPI_ISL_612657|2020 ···················· ···················· ···················· ···················· ···················· ····················
O 1005 hCoV-19/USA/FL-BPHL-1760/2020|EPI_ISL_613868|2020- ···················· ·A·················· ···················· ···················· ···················· ····················

| | | | | | | | | |
|---|---|---|---|---|---|---|---|---|
| O | 1006 hCoV-19/USA/FL-BPHL-1762/2020\|EPI_ISL_613869\|2020- | ···················· | ···················· | ···················· | ···················· | NNN················· | ···················· | 1 |
| O | 1008 hCoV-19/USA/FL-BPHL-1765/2020\|EPI_ISL_613871\|2020- | ···················· | ···················· | ···················· | ···················· | ···················· | ···················· | |
| O | 1011 hCoV-19/USA/FL-BPHL-1769/2020\|EPI_ISL_613874\|2020- | ···················· | ···················· | ···················· | ···················· | ···················· | ···················· | |
| O | 1012 hCoV-19/USA/FL-BPHL-1770/2020\|EPI_ISL_613875\|2020- | ···················· | ···················· | ···················· | ···················· | ···················· | ···················· | |
| O | 1013 hCoV-19/USA/FL-BPHL-1771/2020\|EPI_ISL_613876\|2020- | ···················· | ···················· | ···················· | ···················· | ···················· | ···················· | |
| O | 1014 hCoV-19/USA/FL-BPHL-1772/2020\|EPI_ISL_613877\|2020- | ···················· | ···················· | ···················· | ···················· | ···················· | ···················· | |
| O | 1015 hCoV-19/USA/FL-BPHL-1773/2020\|EPI_ISL_613878\|2020- | ···················· | ···················· | ···················· | ···················· | ···················· | ···················· | |
| O | 1016 hCoV-19/USA/FL-BPHL-1781/2020\|EPI_ISL_613884\|2020- | ···················· | ···················· | ···················· | ···················· | ···················· | ···················· | |
| O | 1017 hCoV-19/USA/FL-BPHL-1782/2020\|EPI_ISL_613885\|2020- | ···················· | ···················· | ···················· | ···················· | ···················· | ···················· | |
| O | 1018 hCoV-19/USA/FL-BPHL-1869/2020\|EPI_ISL_613945\|2020- | ···················· | ···················· | ···················· | ···················· | ···················· | ···················· | |
| O | 1019 hCoV-19/USA/FL-BPHL-1870/2020\|EPI_ISL_613946\|2020- | ···················· | ···················· | ···················· | ···················· | ···················· | ···················· | |
| O | 1020 hCoV-19/USA/FL-BPHL-1871/2020\|EPI_ISL_613947\|2020- | ···················· | ···················· | ···················· | ···················· | ···················· | ···················· | |
| O | 1021 hCoV-19/USA/FL-BPHL-1872/2020\|EPI_ISL_613948\|2020- | ···················· | ···················· | ···················· | ···················· | ···················· | ···················· | |
| O | 1022 hCoV-19/USA/FL-BPHL-1889/2020\|EPI_ISL_613949\|2020- | ···················· | ···················· | ···················· | ···················· | ···················· | ···················· | |
| O | 1023 hCoV-19/Italy/SAR-ASSL-03/2020\|EPI_ISL_613953\|2020 | ···················· | ···················· | ···················· | ···················· | ···················· | ···················· | |
| O | 1024 hCoV-19/Gibraltar/204421181/2020\|EPI_ISL_613958\|20 | ···················· | ···················· | ···················· | ···················· | ·····K·············· | ···················· | |
| O | 1025 hCoV-19/England/MILK-A7D848/2020\|EPI_ISL_606600\|20 | ···················· | ···················· | ···················· | ···················· | ···················· | ···················· | |
| O | 1026 hCoV-19/England/MILK-A7CB5F/2020\|EPI_ISL_606601\|20 | ···················· | ···················· | ···················· | ···················· | ···················· | ···················· | |
| O | 1027 hCoV-19/England/MILK-A7DBE5/2020\|EPI_ISL_606602\|20 | ···················· | ·A·················· | ···················· | ···················· | ···················· | ···A················ | |
| O | 1028 hCoV-19/England/MILK-A7D110/2020\|EPI_ISL_606603\|20 | ···················· | ···················· | ···················· | ···················· | ·····T·············· | ···················· | |
| O | 1029 hCoV-19/England/MILK-A7A81E/2020\|EPI_ISL_606604\|20 | ···················· | ···················· | ···················· | ···················· | ·····T·············· | ···················· | |
| O | 1030 hCoV-19/England/MILK-A795BE/2020\|EPI_ISL_606605\|20 | ···················· | ···················· | ···················· | ···················· | ···················· | ···················· | |
| O | 1031 hCoV-19/England/MILK-A7974F/2020\|EPI_ISL_606606\|20 | ···················· | ···················· | ···················· | ···················· | ···················· | ···················· | |
| O | 1032 hCoV-19/England/MILK-A79D3E/2020\|EPI_ISL_606607\|20 | ···················· | ···················· | ···················· | ···················· | ···················· | ···················· | |
| O | 1033 hCoV-19/England/MILK-A7A41A/2020\|EPI_ISL_606609\|20 | ···················· | ···················· | ···················· | ···················· | ·····T·············· | ···················· | |
| O | 1034 hCoV-19/England/MILK-A79E4A/2020\|EPI_ISL_606610\|20 | ···················· | ···················· | ···················· | ···················· | ···················· | ···················· | |
| O | 1035 hCoV-19/England/MILK-A7CF9F/2020\|EPI_ISL_606611\|20 | ···················· | ···················· | ···················· | ···················· | ···················· | ···················· | |
| O | 1036 hCoV-19/England/MILK-A7A377/2020\|EPI_ISL_606612\|20 | ···················· | ·A·················· | ···················· | ···················· | ···················· | ···················· | |
| O | 1037 hCoV-19/England/MILK-A7D0B9/2020\|EPI_ISL_606613\|20 | ···················· | ···················· | ···················· | ···················· | ···················· | ···················· | |
| O | 1038 hCoV-19/England/MILK-A79FA1/2020\|EPI_ISL_606614\|20 | ···················· | ···················· | ···················· | ···················· | ···················· | ················A··· | |
| O | 1039 hCoV-19/England/MILK-A795EB/2020\|EPI_ISL_606615\|20 | ···················· | ···················· | ···················· | ···················· | ···················· | ···················· | |
| O | 1040 hCoV-19/England/MILK-A7ABF7/2020\|EPI_ISL_606616\|20 | ···················· | ···················· | ···················· | ···················· | ···················· | ···················· | |
| O | 1041 hCoV-19/England/MILK-A79F29/2020\|EPI_ISL_606618\|20 | ···················· | ···················· | ···················· | ···················· | ···················· | ···················· | |
| O | 1042 hCoV-19/England/ALDP-A6F454/2020\|EPI_ISL_607907\|20 | ···················· | ···················· | ···················· | ···················· | ···················· | ···················· | |
| O | 1043 hCoV-19/England/ALDP-A7062D/2020\|EPI_ISL_607908\|20 | ···················· | ···················· | ···················· | ···················· | ···················· | ···················· | |
| O | 1044 hCoV-19/England/ALDP-A6D939/2020\|EPI_ISL_607909\|20 | ···················· | ···················· | ···················· | ···················· | ···················· | ···················· | |
| O | 1045 hCoV-19/England/ALDP-A6E983/2020\|EPI_ISL_607910\|20 | ···················· | ·A·················· | ···················· | ···················· | ···················· | ···A················ | |
| O | 1046 hCoV-19/England/ALDP-A6E23D/2020\|EPI_ISL_607911\|20 | ···················· | ···················· | ···················· | ···················· | ···················· | ···················· | |
| O | 1047 hCoV-19/England/ALDP-A7046F/2020\|EPI_ISL_607912\|20 | ···················· | ···················· | ···················· | ···················· | ···················· | ···················· | |
| O | 1048 hCoV-19/England/ALDP-A6EA53/2020\|EPI_ISL_607913\|20 | ···················· | ···················· | ···················· | ···················· | ···················· | ···················· | |
| O | 1049 hCoV-19/England/ALDP-A6F3EE/2020\|EPI_ISL_607914\|20 | ···················· | ···················· | ···················· | ···················· | ···················· | ···················· | |
| O | 1050 hCoV-19/England/ALDP-A6DE49/2020\|EPI_ISL_607915\|20 | ···················· | ···················· | ···················· | ···················· | ···················· | ···················· | |
| O | 1051 hCoV-19/England/ALDP-A6EFFA/2020\|EPI_ISL_607916\|20 | ···················· | ···················· | ···················· | ···················· | ···················· | ···················· | |
| O | 1052 hCoV-19/England/ALDP-A705C6/2020\|EPI_ISL_607918\|20 | ···················· | ···················· | ···················· | ···················· | ···················· | ···················· | |
| O | 1053 hCoV-19/England/ALDP-A6D887/2020\|EPI_ISL_607920\|20 | ···················· | ···················· | ···················· | ···················· | ···················· | ···················· | |
| O | 1054 hCoV-19/England/ALDP-A7064B/2020\|EPI_ISL_607921\|20 | ···················· | ···················· | ···················· | ···················· | ···················· | ···················· | |
| O | 1055 hCoV-19/England/ALDP-A6E3D0/2020\|EPI_ISL_607922\|20 | ···················· | ···················· | ···················· | ···················· | ···················· | ···················· | |
| O | 1056 hCoV-19/England/ALDP-A6DF55/2020\|EPI_ISL_607923\|20 | ···················· | ···················· | ···················· | ···················· | ···················· | ···················· | |
| O | 1057 hCoV-19/England/ALDP-A6DAEB/2020\|EPI_ISL_607925\|20 | ···················· | ···················· | ···················· | ···················· | ···················· | ···················· | |
| O | 1058 hCoV-19/England/ALDP-A6F0BA/2020\|EPI_ISL_607926\|20 | ···················· | ···················· | ···················· | ···················· | ···················· | ···················· | |
| O | 1059 hCoV-19/England/ALDP-A6F3A2/2020\|EPI_ISL_607927\|20 | ···················· | ···················· | ···················· | ···················· | ···················· | ···················· | |
| O | 1060 hCoV-19/England/ALDP-A6D83C/2020\|EPI_ISL_607928\|20 | ···················· | ···················· | ···················· | ···················· | ···················· | ···················· | |
| O | 1061 hCoV-19/England/ALDP-A6E8C2/2020\|EPI_ISL_607929\|20 | ···················· | ···················· | ···················· | ···················· | ···················· | ···················· | |
| O | 1062 hCoV-19/England/ALDP-A6F5AC/2020\|EPI_ISL_607930\|20 | ···················· | ···················· | ···················· | ···················· | ···················· | ···················· | |
| O | 1063 hCoV-19/England/ALDP-A6E26A/2020\|EPI_ISL_607932\|20 | ···················· | ···················· | ···················· | ···················· | ···················· | ···················· | |
| O | 1064 hCoV-19/England/ALDP-A6DED0/2020\|EPI_ISL_607933\|20 | ···················· | ···················· | ···················· | ···················· | ···················· | ···················· | |
| O | 1065 hCoV-19/England/ALDP-A6E08E/2020\|EPI_ISL_607934\|20 | ···················· | ···················· | ···················· | ···················· | ···················· | ···················· | |
| O | 1066 hCoV-19/England/ALDP-A6E16D/2020\|EPI_ISL_607935\|20 | ···················· | ···················· | ···················· | ···················· | ···················· | ···················· | |
| O | 1067 hCoV-19/England/ALDP-A6F59D/2020\|EPI_ISL_607936\|20 | ···················· | ···················· | ···················· | ···················· | ···················· | ···················· | |
| O | 1068 hCoV-19/England/ALDP-A7060F/2020\|EPI_ISL_607938\|20 | ···················· | ···················· | ···················· | ···················· | ···················· | ···················· | |
| O | 1069 hCoV-19/England/ALDP-A6E8B3/2020\|EPI_ISL_607939\|20 | ···················· | ···················· | ···················· | ···················· | ···················· | ···················· | |
| O | 1070 hCoV-19/England/ALDP-A6E473/2020\|EPI_ISL_607940\|20 | ···················· | ···················· | ···················· | ···················· | ···················· | ···················· | |
| O | 1071 hCoV-19/England/ALDP-A70405/2020\|EPI_ISL_607941\|20 | ···················· | ···················· | ···················· | ···················· | ···················· | ···················· | |
| O | 1072 hCoV-19/England/ALDP-A6EA35/2020\|EPI_ISL_607942\|20 | ···················· | ···················· | ···················· | ···················· | ···················· | ···················· | |
| O | 1073 hCoV-19/England/ALDP-A6F3FD/2020\|EPI_ISL_607943\|20 | ···················· | ···················· | ···················· | ···················· | ···················· | ···················· | |
| O | 1074 hCoV-19/England/ALDP-A6EF45/2020\|EPI_ISL_607944\|20 | ···················· | ···················· | ···················· | ···················· | ···················· | ···················· | |
| O | 1075 hCoV-19/England/ALDP-A6F5D9/2020\|EPI_ISL_607945\|20 | ···················· | ···················· | ···················· | ···················· | ···················· | ···················· | |
| O | 1076 hCoV-19/England/ALDP-A6E3EF/2020\|EPI_ISL_607946\|20 | ···················· | ···················· | ···················· | ···················· | ···················· | ···················· | |
| O | 1077 hCoV-19/England/ALDP-A6DE58/2020\|EPI_ISL_607947\|20 | ···················· | ···················· | ···················· | ···················· | ···················· | ···················· | |
| O | 1078 hCoV-19/England/ALDP-A6F533/2020\|EPI_ISL_607948\|20 | ···················· | ···················· | ···················· | ···················· | ···················· | ···················· | |
| O | 1079 hCoV-19/England/ALDP-A6E90B/2020\|EPI_ISL_607949\|20 | ···················· | ···················· | ···················· | ···················· | ···················· | ···················· | |
| O | 1080 hCoV-19/England/ALDP-A92AED/2020\|EPI_ISL_611129\|20 | ···················· | ···················· | ···················· | ···················· | ···················· | ···················· | |
| O | 1081 hCoV-19/England/ALDP-A948F0/2020\|EPI_ISL_611130\|20 | ···················· | ···················· | ···················· | ···················· | ···················· | ···A················ | |
| O | 1082 hCoV-19/England/ALDP-A93976/2020\|EPI_ISL_611131\|20 | ···················· | ···················· | ···················· | ···················· | ···················· | ···················· | |
| O | 1083 hCoV-19/England/ALDP-A9291D/2020\|EPI_ISL_611132\|20 | ···················· | ···················· | ···················· | ···················· | ···················· | ···················· | |
| O | 1084 hCoV-19/England/ALDP-A93C8C/2020\|EPI_ISL_611133\|20 | ···················· | ···················· | ···················· | ···················· | ···················· | ···················· | |
| O | 1085 hCoV-19/England/ALDP-A938D3/2020\|EPI_ISL_611134\|20 | ···················· | ···················· | ···················· | ···················· | ···················· | ···················· | |
| O | 1086 hCoV-19/England/ALDP-A93651/2020\|EPI_ISL_611135\|20 | ···················· | ·A·················· | ···················· | ···················· | ···················· | ···················· | |
| O | 1087 hCoV-19/England/ALDP-A925CE/2020\|EPI_ISL_611136\|20 | ···················· | ·A·················· | ···················· | ···················· | ···················· | ···A················ | |
| O | 1088 hCoV-19/England/ALDP-A92A74/2020\|EPI_ISL_611138\|20 | ···················· | ···················· | ···················· | ···················· | ···················· | ···················· | |

| | | | | | | | |
|---|---|---|---|---|---|---|---|
| O | 1089 hCoV-19/England/ALDP-A92740/2020\|EPI_ISL_611139\|20 | ···················· | ···················· | ····················· | ···················· | ···················· | ···················· |
| O | 1090 hCoV-19/England/ALDP-A938F1/2020\|EPI_ISL_611143\|20 | ···················· | ···················· | ····················· | ···················· | ···················· | ···················· |
| O | 1091 hCoV-19/England/ALDP-A929A4/2020\|EPI_ISL_611145\|20 | ···················· | ···················· | ····················· | ···················· | ···················· | ···················· |
| O | 1092 hCoV-19/England/ALDP-A93D7A/2020\|EPI_ISL_611146\|20 | ···················· | ·A·················· | ····················· | ···················· | ···················· | ···················· |
| O | 1093 hCoV-19/England/ALDP-A93BE9/2020\|EPI_ISL_611148\|20 | ···················· | ···················· | ····················· | ···················· | ···················· | ···················· |
| O | 1094 hCoV-19/England/ALDP-A93572/2020\|EPI_ISL_611170\|20 | ···················· | ···················· | ····················· | ···················· | ···················· | ···················· |
| O | 1095 hCoV-19/England/ALDP-A92BAE/2020\|EPI_ISL_611171\|20 | ···················· | ···················· | ····················· | ···················· | ···················· | ···················· |
| O | 1096 hCoV-19/England/ALDP-A93A19/2020\|EPI_ISL_611172\|20 | ···················· | ···················· | ····················· | ···················· | ···················· | ···················· |
| O | 1097 hCoV-19/England/ALDP-A948E1/2020\|EPI_ISL_611173\|20 | ···················· | ···················· | ····················· | ···················· | ···················· | ···A················ |
| O | 1098 hCoV-19/England/ALDP-A93563/2020\|EPI_ISL_611174\|20 | ···················· | ·A·················· | ····················· | ···················· | ···················· | ···A················ |
| O | 1099 hCoV-19/England/ALDP-A92B71/2020\|EPI_ISL_611175\|20 | ···················· | ···················· | ····················· | ···················· | ···················· | ···················· |
| O | 1100 hCoV-19/England/ALDP-A92C05/2020\|EPI_ISL_611176\|20 | ···················· | ···················· | ····················· | ···················· | ···················· | ···················· |
| O | 1101 hCoV-19/England/ALDP-A93A64/2020\|EPI_ISL_611177\|20 | ···················· | ···················· | ····················· | ···················· | ···················· | ···················· |
| O | 1102 hCoV-19/England/ALDP-A92D3F/2020\|EPI_ISL_611178\|20 | ···················· | ···················· | ····················· | ···················· | ···················· | ···················· |
| O | 1103 hCoV-19/England/ALDP-A93B61/2020\|EPI_ISL_611179\|20 | ···················· | ···················· | ····················· | ···················· | ···················· | ···················· |
| O | 1104 hCoV-19/England/ALDP-A93A73/2020\|EPI_ISL_611181\|20 | ···················· | ···················· | ····················· | ···················· | ···················· | ···················· |
| O | 1105 hCoV-19/England/ALDP-A93DF2/2020\|EPI_ISL_611182\|20 | ···················· | ···················· | ····················· | ···················· | ···················· | ···················· |
| O | 1106 hCoV-19/England/ALDP-A936F7/2020\|EPI_ISL_611183\|20 | ···················· | ···················· | ····················· | ···················· | ···················· | ···················· |
| O | 1107 hCoV-19/England/ALDP-A93CE6/2020\|EPI_ISL_611184\|20 | ···················· | ···················· | ····················· | ···················· | ···················· | ···················· |
| O | 1108 hCoV-19/England/ALDP-A92A65/2020\|EPI_ISL_611185\|20 | ···················· | ···················· | ····················· | ···················· | ···················· | ···················· |
| O | 1109 hCoV-19/England/ALDP-A947F3/2020\|EPI_ISL_611186\|20 | ···················· | ···················· | ····················· | ···················· | ···················· | ···················· |
| O | 1110 hCoV-19/England/ALDP-A92E4B/2020\|EPI_ISL_611188\|20 | ···················· | ···················· | ····················· | ···················· | ···················· | ···················· |
| O | 1111 hCoV-19/England/ALDP-A92C6F/2020\|EPI_ISL_611189\|20 | ···················· | ···················· | ····················· | ···················· | ···················· | ···················· |
| O | 1112 hCoV-19/England/ALDP-A94034/2020\|EPI_ISL_611191\|20 | ···················· | ···················· | ····················· | ···················· | ···················· | ···················· |
| O | 1113 hCoV-19/England/ALDP-A92643/2020\|EPI_ISL_611192\|20 | ···················· | ···················· | ····················· | ···················· | ···················· | ···················· |
| O | 1114 hCoV-19/England/ALDP-A93EA4/2020\|EPI_ISL_611193\|20 | ···················· | ···················· | ····················· | ···················· | ···················· | ···················· |
| O | 1115 hCoV-19/England/ALDP-A92ED2/2020\|EPI_ISL_611194\|20 | ···················· | ···················· | ····················· | ···················· | ···················· | ···················· |
| O | 1116 hCoV-19/England/ALDP-A92D20/2020\|EPI_ISL_611195\|20 | ···················· | ···················· | ····················· | ···················· | ···················· | ···················· |
| O | 1117 hCoV-19/England/ALDP-A96EB0/2020\|EPI_ISL_611198\|20 | ···················· | ···················· | ····················· | ···················· | ···················· | ···················· |
| O | 1118 hCoV-19/England/ALDP-A93FDE/2020\|EPI_ISL_611199\|20 | ···················· | ···················· | ····················· | ···················· | ···················· | ···················· |
| O | 1119 hCoV-19/England/ALDP-A94016/2020\|EPI_ISL_611200\|20 | ···················· | ···················· | ····················· | ···················· | ···················· | ···················· |
| O | 1120 hCoV-19/England/ALDP-A936BB/2020\|EPI_ISL_611201\|20 | ···················· | ···················· | ····················· | ···················· | ···················· | ···················· |
| O | 1121 hCoV-19/England/ALDP-A9491B/2020\|EPI_ISL_611202\|20 | ···················· | ···················· | ····················· | ···················· | ···················· | ···················· |
| O | 1122 hCoV-19/England/ALDP-A9285C/2020\|EPI_ISL_611203\|20 | ···················· | ···················· | ····················· | ···················· | ···················· | ···················· |
| O | 1123 hCoV-19/England/ALDP-A93D6B/2020\|EPI_ISL_611205\|20 | ···················· | ···················· | ····················· | ···················· | ···················· | ···················· |
| O | 1124 hCoV-19/England/ALDP-A93888/2020\|EPI_ISL_611217\|20 | ···················· | ···················· | ····················· | ···················· | ···················· | ···················· |
| O | 1125 hCoV-19/England/ALDP-A92CD8/2020\|EPI_ISL_611218\|20 | ···················· | ···················· | ····················· | ···················· | ···················· | ···················· |
| O | 1126 hCoV-19/England/ALDP-A935CD/2020\|EPI_ISL_611219\|20 | ···················· | ···················· | ····················· | ···················· | ···················· | ···················· |
| O | 1127 hCoV-19/England/ALDP-A93B16/2020\|EPI_ISL_611222\|20 | ···················· | ···················· | ····················· | ···················· | ···················· | ···················· |
| O | 1128 hCoV-19/England/ALDP-A93A0A/2020\|EPI_ISL_611223\|20 | ···················· | ···················· | ····················· | ···················· | ···················· | ···················· |
| O | 1129 hCoV-19/England/ALDP-A93590/2020\|EPI_ISL_611224\|20 | ···················· | ···················· | ····················· | ···················· | ···················· | ···················· |
| O | 1130 hCoV-19/England/ALDP-A92BF9/2020\|EPI_ISL_611226\|20 | ···················· | ···················· | ····················· | ···················· | ···················· | ···················· |
| O | 1131 hCoV-19/England/ALDP-A925DD/2020\|EPI_ISL_611227\|20 | ···················· | ···················· | ····················· | ···················· | ···················· | ···················· |
| O | 1132 hCoV-19/England/ALDP-A927C8/2020\|EPI_ISL_611228\|20 | ···················· | ···················· | ····················· | ···················· | ···················· | ···················· |
| O | 1133 hCoV-19/England/ALDP-A9293B/2020\|EPI_ISL_611229\|20 | ···················· | ···················· | ····················· | ···················· | ···················· | ···················· |
| O | 1134 hCoV-19/England/ALDP-A938C4/2020\|EPI_ISL_611230\|20 | ···················· | ···················· | ····················· | ···················· | ···················· | ···················· |
| O | 1135 hCoV-19/England/ALDP-A948D2/2020\|EPI_ISL_611231\|20 | ···················· | ···················· | ····················· | ···················· | ···················· | ···················· |
| O | 1136 hCoV-19/England/ALDP-A9367F/2020\|EPI_ISL_611232\|20 | ···················· | ···················· | ····················· | ···················· | ···················· | ···················· |
| O | 1137 hCoV-19/England/ALDP-A92591/2020\|EPI_ISL_611234\|20 | ···················· | ···················· | ····················· | ···················· | ···················· | ···················· |
| O | 1138 hCoV-19/England/ALDP-A93615/2020\|EPI_ISL_611235\|20 | ···················· | ···················· | ····················· | ···················· | ···················· | ···················· |
| O | 1139 hCoV-19/England/ALDP-A92573/2020\|EPI_ISL_611236\|20 | ···················· | ···················· | ····················· | ···················· | ···················· | ···················· |
| O | 1140 hCoV-19/England/ALDP-A93E95/2020\|EPI_ISL_611238\|20 | ···················· | ···················· | ····················· | ···················· | ···················· | ···················· |
| O | 1141 hCoV-19/England/ALDP-A93A28/2020\|EPI_ISL_611240\|20 | ···················· | ···················· | ····················· | ···················· | ···················· | ···················· |
| O | 1142 hCoV-19/England/ALDP-A92555/2020\|EPI_ISL_611241\|20 | ···················· | ···················· | ····················· | ···················· | ···················· | ···················· |
| O | 1143 hCoV-19/England/ALDP-A92EE1/2020\|EPI_ISL_611243\|20 | ···················· | ···················· | ····················· | ···················· | ···················· | ···················· |
| O | 1144 hCoV-19/England/ALDP-A935BE/2020\|EPI_ISL_611244\|20 | ···················· | ···················· | ····················· | ···················· | ···················· | ···················· |
| O | 1145 hCoV-19/England/ALDP-A92DC6/2020\|EPI_ISL_611246\|20 | ···················· | ···················· | ····················· | ···················· | ···················· | ···················· |
| O | 1146 hCoV-19/England/ALDP-A92ACF/2020\|EPI_ISL_611247\|20 | ···················· | ···················· | ····················· | ···················· | ···················· | ···················· |
| O | 1147 hCoV-19/England/ALDP-A92B9F/2020\|EPI_ISL_611249\|20 | ···················· | ···················· | ····················· | ···················· | ···················· | ···················· |
| O | 1148 hCoV-19/England/MILK-A18DC8/2020\|EPI_ISL_612460\|20 | ···················· | ···················· | ····················· | ···················· | ···················· | ···················· |
| O | 1149 hCoV-19/England/MILK-A18DD7/2020\|EPI_ISL_612461\|20 | ···················· | ···················· | ····················· | ···················· | ···················· | ···················· |
| O | 1150 hCoV-19/England/MILK-A18DE6/2020\|EPI_ISL_612462\|20 | ···················· | ···················· | ····················· | ···················· | ···················· | ···················· |
| O | 1151 hCoV-19/England/MILK-A18DF5/2020\|EPI_ISL_612463\|20 | ···················· | ···················· | ····················· | ···················· | ···················· | ···················· |
| O | 1152 hCoV-19/England/MILK-A18E10/2020\|EPI_ISL_612464\|20 | ···················· | ·R·················· | ····················· | ···················· | ···················· | ···R················ |
| O | 1153 hCoV-19/England/MILK-A18E2F/2020\|EPI_ISL_612465\|20 | ···················· | ···················· | ····················· | ···················· | ···················· | ···················· |
| O | 1154 hCoV-19/England/MILK-A18E3E/2020\|EPI_ISL_612466\|20 | ···················· | ···················· | ····················· | ···················· | ······T············· | ···················· |
| O | 1155 hCoV-19/England/MILK-A18E4D/2020\|EPI_ISL_612467\|20 | ···················· | ···················· | ····················· | ···················· | ······T············· | ···················· |
| O | 1156 hCoV-19/England/MILK-A18E5C/2020\|EPI_ISL_612468\|20 | ···················· | ···················· | ····················· | ···················· | ······T············· | ···················· |
| O | 1157 hCoV-19/England/MILK-A18E6B/2020\|EPI_ISL_612469\|20 | ···················· | ···················· | ····················· | ···················· | ······T············· | ···················· |
| O | 1158 hCoV-19/England/MILK-A18EF2/2020\|EPI_ISL_612474\|20 | ···················· | ···················· | ····················· | ···················· | ···················· | ···················· |
| O | 1159 hCoV-19/England/MILK-A18F0E/2020\|EPI_ISL_612475\|20 | ···················· | ·A·················· | ····················· | ···················· | ···················· | ···A················ |
| O | 1160 hCoV-19/England/MILK-A18F86/2020\|EPI_ISL_612476\|20 | ···················· | ···················· | ····················· | ···················· | ···················· | ···················· |
| O | 1161 hCoV-19/England/MILK-A18F95/2020\|EPI_ISL_612477\|20 | ···················· | ···················· | ····················· | ···················· | ······T············· | ···················· |
| O | 1162 hCoV-19/England/MILK-A192F5/2020\|EPI_ISL_612500\|20 | ···················· | ···················· | ····················· | ···················· | ···················· | ···················· |
| O | 1163 hCoV-19/England/MILK-A19301/2020\|EPI_ISL_612501\|20 | ···················· | ···················· | ····················· | ···················· | ···················· | ···················· |
| O | 1164 hCoV-19/England/MILK-A19310/2020\|EPI_ISL_612502\|20 | ···················· | ···················· | ····················· | ···················· | ···················· | ···················· |
| O | 1165 hCoV-19/England/MILK-A1932F/2020\|EPI_ISL_612503\|20 | ···················· | ···················· | ····················· | ···················· | ···················· | ···················· |
| O | 1166 hCoV-19/England/MILK-A1933E/2020\|EPI_ISL_612504\|20 | ···················· | ···················· | ····················· | ···················· | ···················· | ···················· |
| O | 1167 hCoV-19/England/MILK-A1935C/2020\|EPI_ISL_612505\|20 | ···················· | ···················· | ····················· | ···················· | ···················· | ···················· |
| O | 1168 hCoV-19/England/MILK-A1936B/2020\|EPI_ISL_612506\|20 | ···················· | ···················· | ····················· | ···················· | ···················· | ···················· |

| | | | | | | | | | |
|---|---|---|---|---|---|---|---|---|---|
| O | 1169 | hCoV-19/England/MILK-A1937A/2020\|EPI_ISL_612507\|20 | ···················· | ···················· | ···················· | ···················· | ···················· | ···················· | |
| O | 1170 | hCoV-19/England/MILK-A19398/2020\|EPI_ISL_612508\|20 | ···················· | ·A·················· | ···················· | ···················· | ·········T·········· | ···················· | |
| O | 1172 | hCoV-19/England/MILK-A1942C/2020\|EPI_ISL_612510\|20 | ···················· | ···················· | ···················· | ···················· | ···················· | ···················· | |
| O | 1173 | hCoV-19/England/MILK-A194D1/2020\|EPI_ISL_612511\|20 | ···················· | ···················· | ···················· | ···················· | NNN················· | ···················· | 1 |
| O | 1174 | hCoV-19/England/MILK-A194E0/2020\|EPI_ISL_612512\|20 | ···················· | ···················· | ···················· | ···················· | ···················· | ···················· | |
| O | 1175 | hCoV-19/England/MILK-A194FF/2020\|EPI_ISL_612513\|20 | ···················· | ···················· | ···················· | ···················· | ···················· | ···················· | |
| O | 1176 | hCoV-19/England/MILK-A1951A/2020\|EPI_ISL_612514\|20 | ···················· | ···················· | ···················· | ···················· | ···················· | ···················· | |
| O | 1177 | hCoV-19/England/MILK-A19529/2020\|EPI_ISL_612515\|20 | ···················· | ···················· | ···················· | ···················· | ···················· | ···················· | |
| O | 1179 | hCoV-19/England/MILK-A19556/2020\|EPI_ISL_612517\|20 | ···················· | ···················· | ···················· | ···················· | ···················· | ···················· | |
| O | 1180 | hCoV-19/Wales/MILK-A19583/2020\|EPI_ISL_612518\|2020 | ···················· | ···················· | ···················· | ···················· | ···················· | ···················· | |
| O | 1181 | hCoV-19/Wales/MILK-A19592/2020\|EPI_ISL_612519\|2020 | ···················· | ···················· | ···················· | ···················· | ···················· | ···················· | |
| O | 1182 | hCoV-19/Wales/MILK-A195B0/2020\|EPI_ISL_612520\|2020 | ···················· | ···················· | ···················· | ···················· | ···················· | ···················· | |
| O | 1183 | hCoV-19/Wales/MILK-A195CF/2020\|EPI_ISL_612521\|2020 | ···················· | ···················· | ···················· | ···················· | ···················· | ···················· | |
| O | 1184 | hCoV-19/Wales/MILK-A195DE/2020\|EPI_ISL_612522\|2020 | ···················· | ···················· | ···················· | ···················· | ···················· | ···················· | |
| O | 1185 | hCoV-19/Wales/MILK-A22815/2020\|EPI_ISL_612523\|2020 | ···················· | ···················· | ···················· | ···················· | ···················· | ···················· | |
| O | 1186 | hCoV-19/England/MILK-A22833/2020\|EPI_ISL_612524\|20 | ···················· | ···················· | ···················· | ···················· | ···················· | ···················· | |
| O | 1187 | hCoV-19/England/MILK-A2289D/2020\|EPI_ISL_612525\|20 | ···················· | ···················· | ···················· | ···················· | ···················· | ···················· | |
| O | 1188 | hCoV-19/England/MILK-A228E8/2020\|EPI_ISL_612526\|20 | ···················· | ···················· | ···················· | ···················· | ···················· | ···················· | |
| O | 1189 | hCoV-19/England/MILK-A228F7/2020\|EPI_ISL_612527\|20 | ···················· | ···················· | ···················· | ···················· | ···················· | ···················· | |
| O | 1190 | hCoV-19/England/MILK-A22903/2020\|EPI_ISL_612528\|20 | ···················· | ···················· | ···················· | ···················· | ···················· | ···················· | |
| O | 1191 | hCoV-19/England/MILK-A2294F/2020\|EPI_ISL_612529\|20 | ···················· | ···················· | ···················· | ···················· | ···················· | ···················· | |
| O | 1192 | hCoV-19/England/MILK-A2295E/2020\|EPI_ISL_612530\|20 | ···················· | ···················· | ···················· | ···················· | ···················· | ···················· | |
| O | 1193 | hCoV-19/England/MILK-A2296D/2020\|EPI_ISL_612531\|20 | ···················· | ···················· | ···················· | ···················· | ···················· | ···················· | |
| O | 1194 | hCoV-19/England/MILK-A2297C/2020\|EPI_ISL_612532\|20 | ···················· | ···················· | ···················· | ···················· | ···················· | ···················· | |
| O | 1196 | hCoV-19/England/MILK-A229C7/2020\|EPI_ISL_612534\|20 | ···················· | ···················· | ···················· | ···················· | ···················· | ···················· | |
| O | 1197 | hCoV-19/England/MILK-A229D6/2020\|EPI_ISL_612535\|20 | ···················· | ···················· | ···················· | ···················· | ···················· | ···················· | |
| O | 1198 | hCoV-19/England/MILK-A229E5/2020\|EPI_ISL_612536\|20 | ···················· | ·····A·············· | ···················· | ···················· | ···················· | ···················· | |
| O | 1199 | hCoV-19/Northern Ireland/NIRE-108F55/2020\|EPI_ISL_ | ···················· | ···················· | ···················· | ···················· | ···················· | ···················· | |
| O | 1200 | hCoV-19/England/NORT-1B610D2/2020\|EPI_ISL_612549\|2 | ···················· | ·A·················· | ···················· | ···················· | ···················· | ···················· | |
| O | 1201 | hCoV-19/England/NORT-1B6BECF/2020\|EPI_ISL_612550\|2 | ···················· | ·A·················· | ···················· | ···················· | ···················· | ···················· | |
| O | 1202 | hCoV-19/England/NORT-1B6BEDE/2020\|EPI_ISL_612551\|2 | ···················· | ···················· | ···················· | ···················· | ···················· | ···················· | |
| O | 1203 | hCoV-19/England/NORT-1B6BF08/2020\|EPI_ISL_612552\|2 | ···················· | ···················· | ···················· | ···················· | ···················· | ···················· | |
| O | 1204 | hCoV-19/England/NORT-1B6BF35/2020\|EPI_ISL_612554\|2 | ···················· | ···················· | ···················· | ···················· | ···················· | ···················· | |
| O | 1205 | hCoV-19/England/NORT-1B6BF44/2020\|EPI_ISL_612555\|2 | ···················· | ···················· | ···················· | ···················· | ···················· | ···················· | |
| O | 1206 | hCoV-19/England/NORT-1B6BF9F/2020\|EPI_ISL_612556\|2 | ···················· | ···················· | ···················· | ···················· | ···················· | ···················· | |
| O | 1207 | hCoV-19/England/NORT-1B6C040/2020\|EPI_ISL_612557\|2 | ···················· | ···················· | ···················· | ···················· | ···················· | ···················· | |
| O | 1208 | hCoV-19/England/NORT-1B6C07D/2020\|EPI_ISL_612558\|2 | ···················· | ···················· | ···················· | ···················· | ···················· | ···················· | |
| O | 1209 | hCoV-19/England/NORT-1B6C08C/2020\|EPI_ISL_612559\|2 | ···················· | ···················· | ···················· | ···················· | ···················· | ···················· | |
| O | 1210 | hCoV-19/England/NORT-1B6C0B9/2020\|EPI_ISL_612560\|2 | ···················· | ···················· | ···················· | ···················· | ···················· | ···················· | |
| O | 1211 | hCoV-19/England/NORT-1B6C0C8/2020\|EPI_ISL_612561\|2 | ···················· | ···················· | ···················· | ···················· | ···················· | ···················· | |
| O | 1212 | hCoV-19/England/NORT-1B6C0D7/2020\|EPI_ISL_612562\|2 | ···················· | ···················· | ···················· | ···················· | ···················· | ···················· | |
| O | 1213 | hCoV-19/England/NORT-1B6C0E6/2020\|EPI_ISL_612563\|2 | ···················· | ·A·················· | ···················· | ···················· | ···················· | ···A················ | |
| O | 1214 | hCoV-19/England/NORT-1B6C101/2020\|EPI_ISL_612564\|2 | ···················· | ···················· | ···················· | ···················· | ···················· | ···················· | |
| O | 1215 | hCoV-19/England/NORT-1B6C110/2020\|EPI_ISL_612565\|2 | ···················· | ···················· | ···················· | ···················· | ···················· | ···················· | |
| O | 1216 | hCoV-19/England/NORT-1B6C14D/2020\|EPI_ISL_612566\|2 | ···················· | ···················· | ···················· | ···················· | ···················· | ···················· | |
| O | 1217 | hCoV-19/England/NORT-2A7F93/2020\|EPI_ISL_612567\|20 | ···················· | ·A·················· | ···················· | ···················· | ···················· | ···················· | |
| O | 1218 | hCoV-19/England/NORT-2A7FA2/2020\|EPI_ISL_612568\|20 | ···················· | ···················· | ···················· | ···················· | ···················· | ···················· | |
| O | 1219 | hCoV-19/England/NORT-2A7FEE/2020\|EPI_ISL_612569\|20 | ···················· | ···················· | ···················· | ···················· | ···················· | ···················· | |
| O | 1220 | hCoV-19/England/NORT-2A80AE/2020\|EPI_ISL_612570\|20 | ···················· | ···················· | ···················· | ···················· | ···················· | ···················· | |
| O | 1221 | hCoV-19/England/NOTT-11677A/2020\|EPI_ISL_612571\|20 | ···················· | ···················· | ···················· | ···················· | ···················· | ···················· | |
| O | 1222 | hCoV-19/England/NOTT-11690B/2020\|EPI_ISL_612572\|20 | ···················· | ···················· | ···················· | ···················· | ···················· | ···················· | |
| O | 1223 | hCoV-19/England/NOTT-116BE7/2020\|EPI_ISL_612573\|20 | ···················· | ···················· | ···················· | ···················· | ···················· | ···················· | |
| O | 1224 | hCoV-19/England/NOTT-116D69/2020\|EPI_ISL_612574\|20 | ···················· | ···················· | ···················· | ···················· | ···················· | ···················· | |
| O | 1225 | hCoV-19/England/NOTT-116F63/2020\|EPI_ISL_612575\|20 | ···················· | ···················· | ···················· | ···················· | ···················· | ···················· | |
| O | 1226 | hCoV-19/England/NOTT-116F72/2020\|EPI_ISL_612576\|20 | ···················· | ···················· | ···················· | ···················· | ···················· | ···················· | |
| O | 1227 | hCoV-19/England/NOTT-117630/2020\|EPI_ISL_612577\|20 | ···················· | ···················· | ···················· | ···················· | ···················· | ···················· | |
| O | 1228 | hCoV-19/England/NOTT-11767C/2020\|EPI_ISL_612578\|20 | ···················· | ···················· | ···················· | ···················· | ···················· | ···················· | |
| O | 1229 | hCoV-19/England/NOTT-11768B/2020\|EPI_ISL_612579\|20 | ···················· | ···················· | ···················· | ···················· | ···················· | ···················· | |
| O | 1230 | hCoV-19/USA/FL-BPHL-1897/2020\|EPI_ISL_613829\|2020- | ···················· | ···················· | ···················· | ···················· | ···················· | ···················· | |
| O | 1231 | hCoV-19/USA/FL-BPHL-1929/2020\|EPI_ISL_613830\|2020- | ···················· | ···················· | ···················· | ···················· | ·NN················· | ···················· | |
| O | 1232 | hCoV-19/USA/FL-BPHL-1903/2020\|EPI_ISL_613831\|2020- | ···················· | ···················· | ···················· | ···················· | ···················· | ···················· | |
| O | 1235 | hCoV-19/USA/FL-BPHL-1910/2020\|EPI_ISL_613834\|2020- | ···················· | ···················· | ···················· | ···················· | ···················· | ···················· | |
| O | 1236 | hCoV-19/USA/FL-BPHL-1705/2020\|EPI_ISL_613837\|2020- | ···················· | ···················· | ···················· | ···················· | ···················· | ···················· | |
| O | 1237 | hCoV-19/USA/FL-BPHL-1708/2020\|EPI_ISL_613843\|2020- | ···················· | ·A·················· | ···················· | ···················· | ···················· | ···················· | |
| O | 1240 | hCoV-19/USA/FL-BPHL-1714/2020\|EPI_ISL_613846\|2020- | ···················· | ···················· | ···················· | ···················· | ···················· | ···················· | |
| O | 1241 | hCoV-19/USA/FL-BPHL-1715/2020\|EPI_ISL_613847\|2020- | ···················· | ···················· | ···················· | ···················· | ···················· | ···················· | |
| O | 1243 | hCoV-19/USA/FL-BPHL-1719/2020\|EPI_ISL_613849\|2020- | ···················· | ············A······· | ···················· | ···················· | ···················· | ···················· | |
| O | 1244 | hCoV-19/USA/FL-BPHL-1723/2020\|EPI_ISL_613850\|2020- | ···················· | ···················· | ···················· | ···················· | ···················· | ···················· | |
| O | 1245 | hCoV-19/USA/FL-BPHL-1724/2020\|EPI_ISL_613851\|2020- | ···················· | ···················· | ···················· | ···················· | ···················· | ···················· | |
| O | 1246 | hCoV-19/USA/FL-BPHL-1725/2020\|EPI_ISL_613852\|2020- | ···················· | ···················· | ···················· | ···················· | ···················· | ···················· | |
| O | 1247 | hCoV-19/USA/FL-BPHL-1727/2020\|EPI_ISL_613853\|2020- | ···················· | ···················· | ···················· | ···················· | ···················· | ···················· | |
| O | 1248 | hCoV-19/USA/FL-BPHL-1728/2020\|EPI_ISL_613854\|2020- | ···················· | ···················· | ···················· | ···················· | ···················· | ···················· | |
| O | 1249 | hCoV-19/USA/FL-BPHL-1729/2020\|EPI_ISL_613855\|2020- | ···················· | ·A·················· | ···················· | ···················· | NNN················· | ···················· | 1 |
| O | 1250 | hCoV-19/USA/FL-BPHL-1734/2020\|EPI_ISL_613856\|2020- | ···················· | ············A······· | ···················· | ···················· | ···················· | ···················· | |
| O | 1251 | hCoV-19/USA/FL-BPHL-1736/2020\|EPI_ISL_613857\|2020- | ···················· | ···················· | ···················· | ···················· | ···················· | ···················· | |
| O | 1252 | hCoV-19/USA/FL-BPHL-1738/2020\|EPI_ISL_613858\|2020- | ···················· | ···················· | ···················· | ···················· | ···················· | ···················· | |
| O | 1253 | hCoV-19/USA/FL-BPHL-1741/2020\|EPI_ISL_613859\|2020- | ···················· | ···················· | ···················· | ···················· | ···················· | ···················· | |
| O | 1255 | hCoV-19/USA/FL-BPHL-1743/2020\|EPI_ISL_613861\|2020- | ···················· | ···················· | ···················· | ···················· | ···················· | ···················· | |
| O | 1256 | hCoV-19/USA/FL-BPHL-1744/2020\|EPI_ISL_613862\|2020- | ···················· | ···················· | ···················· | ···················· | ···················· | ···················· | |
| O | 1257 | hCoV-19/USA/FL-BPHL-1746/2020\|EPI_ISL_613863\|2020- | ···················· | ···················· | ···················· | ···················· | ···················· | ···················· | |

| | | | | | | | | |
|---|---|---|---|---|---|---|---|---|
| O | 1258 hCoV-19/USA/FL-BPHL-1748/2020\|EPI_ISL_613864\|2020- | ···················· | ···················· | ···················· | ···················· | ···················· | ···················· | |
| O | 1259 hCoV-19/USA/FL-BPHL-1750/2020\|EPI_ISL_613865\|2020- | ···················· | ·A·················· | ···················· | ···················· | ···················· | ···················· | |
| O | 1260 hCoV-19/USA/FL-BPHL-1758/2020\|EPI_ISL_613866\|2020- | ···················· | ·············A······ | ···················· | ···················· | ···················· | ···················· | |
| O | 1261 hCoV-19/USA/FL-BPHL-1759/2020\|EPI_ISL_613867\|2020- | ···················· | ···················· | ···················· | ···················· | ···················· | ···················· | |
| O | 1262 hCoV-19/USA/FL-BPHL-1774/2020\|EPI_ISL_613879\|2020- | ···················· | ···················· | ···················· | ···················· | ·NNN················ | ···················· | 1 |
| O | 1263 hCoV-19/USA/FL-BPHL-1775/2020\|EPI_ISL_613880\|2020- | ···················· | ···················· | ···················· | ···················· | ···················· | ···················· | |
| O | 1264 hCoV-19/USA/FL-BPHL-1778/2020\|EPI_ISL_613881\|2020- | ···················· | ···················· | ···················· | ···················· | ···················· | ···················· | |
| O | 1265 hCoV-19/USA/FL-BPHL-1779/2020\|EPI_ISL_613882\|2020- | ···················· | ···················· | ···················· | ···················· | ···················· | ···················· | |
| O | 1266 hCoV-19/USA/FL-BPHL-1780/2020\|EPI_ISL_613883\|2020- | ···················· | ···················· | ···················· | ···················· | ···················· | ···················· | |
| O | 1267 hCoV-19/USA/FL-BPHL-1783/2020\|EPI_ISL_613886\|2020- | ···················· | ···················· | ···················· | ···················· | ···················· | ···················· | |
| O | 1268 hCoV-19/USA/FL-BPHL-1784/2020\|EPI_ISL_613887\|2020- | ···················· | ···················· | ···················· | ···················· | ···················· | ···················· | |
| O | 1269 hCoV-19/USA/FL-BPHL-1786/2020\|EPI_ISL_613888\|2020- | ···················· | ···················· | ···················· | ···················· | ···················· | ···················· | |
| O | 1270 hCoV-19/USA/FL-BPHL-1787/2020\|EPI_ISL_613889\|2020- | ···················· | ···················· | ···················· | ···················· | ···················· | ···················· | |
| O | 1271 hCoV-19/USA/FL-BPHL-1788/2020\|EPI_ISL_613890\|2020- | ···················· | ·A·················· | ···················· | ···················· | ···················· | ···················· | |
| O | 1272 hCoV-19/USA/FL-BPHL-1789/2020\|EPI_ISL_613891\|2020- | ···················· | ···················· | ···················· | ···················· | ···················· | ···················· | |
| O | 1274 hCoV-19/USA/FL-BPHL-1791/2020\|EPI_ISL_613893\|2020- | ···················· | ···················· | ···················· | ···················· | ···················· | ···················· | |
| O | 1275 hCoV-19/USA/FL-BPHL-1792/2020\|EPI_ISL_613894\|2020- | ···················· | ···················· | ···················· | ···················· | ···················· | ···················· | |
| O | 1276 hCoV-19/USA/FL-BPHL-1796/2020\|EPI_ISL_613895\|2020- | ···················· | ···················· | ···················· | ···················· | ···················· | ···················· | |
| O | 1277 hCoV-19/USA/FL-BPHL-1797/2020\|EPI_ISL_613896\|2020- | ···················· | ···················· | ···················· | ···················· | ···················· | ···················· | |
| O | 1278 hCoV-19/USA/FL-BPHL-1798/2020\|EPI_ISL_613897\|2020- | ···················· | ···················· | ···················· | ···················· | ···················· | ···················· | |
| O | 1279 hCoV-19/USA/FL-BPHL-1799/2020\|EPI_ISL_613898\|2020- | ···················· | ···················· | ···················· | ···················· | ···················· | ···················· | |
| O | 1281 hCoV-19/England/MILK-A7C3FD/2020\|EPI_ISL_606580\|20 | ···················· | ···················· | ···················· | ···················· | ···················· | ···················· | |
| O | 1282 hCoV-19/England/MILK-A7A896/2020\|EPI_ISL_606581\|20 | ···················· | ···················· | ···················· | ···················· | ···················· | ···················· | |
| O | 1283 hCoV-19/England/MILK-A7D8C0/2020\|EPI_ISL_606582\|20 | ···················· | ···················· | ···················· | ···················· | ···················· | ···················· | |
| O | 1284 hCoV-19/England/MILK-A7A984/2020\|EPI_ISL_606583\|20 | ···················· | ···················· | ···················· | ···················· | ···················· | ···················· | |
| O | 1285 hCoV-19/England/MILK-A7D7B4/2020\|EPI_ISL_606584\|20 | ···················· | ···················· | ···················· | ···················· | ···················· | ···················· | |
| O | 1286 hCoV-19/England/MILK-A7A6D8/2020\|EPI_ISL_606585\|20 | ···················· | ···················· | ···················· | ···················· | ···················· | ···················· | |
| O | 1287 hCoV-19/England/MILK-A7D3DE/2020\|EPI_ISL_606586\|20 | ···················· | ···················· | ···················· | ···················· | ···················· | ···················· | |
| O | 1288 hCoV-19/England/MILK-A7A9DF/2020\|EPI_ISL_606587\|20 | ···················· | ···················· | ···················· | ···················· | ···················· | ···················· | |
| O | 1289 hCoV-19/England/MILK-A7A6BA/2020\|EPI_ISL_606589\|20 | ···················· | ···················· | ···················· | ···················· | ···················· | ···················· | |
| O | 1290 hCoV-19/England/MILK-A7D15C/2020\|EPI_ISL_606590\|20 | ···················· | ···················· | ···················· | ···················· | ···················· | ···················· | |
| O | 1291 hCoV-19/England/MILK-A7A966/2020\|EPI_ISL_606591\|20 | ···················· | ·A·······A·········· | ···················· | ···················· | ···················· | ···················· | |
| O | 1292 hCoV-19/England/MILK-A7A939/2020\|EPI_ISL_606592\|20 | ···················· | ···················· | ···················· | ···················· | ···················· | ···················· | |
| O | 1293 hCoV-19/England/MILK-A7D866/2020\|EPI_ISL_606593\|20 | ···················· | ···················· | ···················· | ···················· | ···················· | ···················· | |
| O | 1294 hCoV-19/England/MILK-A7A75D/2020\|EPI_ISL_606594\|20 | ···················· | ···················· | ···················· | ···················· | ···················· | ···················· | |
| O | 1295 hCoV-19/England/MILK-A7A7F3/2020\|EPI_ISL_606595\|20 | ···················· | ···················· | ···················· | ···················· | ···················· | ···················· | |
| O | 1296 hCoV-19/England/MILK-A7CD68/2020\|EPI_ISL_606596\|20 | ···················· | ···················· | ···················· | ···················· | ···················· | ···················· | |
| O | 1297 hCoV-19/England/MILK-A7C06F/2020\|EPI_ISL_606597\|20 | ···················· | ···················· | ···················· | ···················· | ···················· | ···················· | |
| O | 1298 hCoV-19/England/MILK-A7A85A/2020\|EPI_ISL_606599\|20 | ···················· | ···················· | ···················· | ···················· | ···················· | ···················· | |
| O | 1299 hCoV-19/England/MILK-A79F1A/2020\|EPI_ISL_606619\|20 | ···················· | ···················· | ···················· | ···················· | ···················· | ···················· | |
| O | 1300 hCoV-19/England/MILK-A7A4FC/2020\|EPI_ISL_606620\|20 | ···················· | ···················· | ···················· | ···················· | ···················· | ···················· | |
| O | 1301 hCoV-19/England/MILK-A7A0BC/2020\|EPI_ISL_606621\|20 | ···················· | ···················· | ···················· | ···················· | ······T············· | ···················· | |
| O | 1302 hCoV-19/England/MILK-A79A46/2020\|EPI_ISL_606623\|20 | ···················· | ···················· | ···················· | ···················· | ···················· | ···················· | |
| O | 1303 hCoV-19/England/MILK-A79EA4/2020\|EPI_ISL_606625\|20 | ···················· | ···················· | ···················· | ···················· | ···················· | ···················· | |
| O | 1304 hCoV-19/England/MILK-A79DE3/2020\|EPI_ISL_606626\|20 | ···················· | ···················· | ···················· | ···················· | ···················· | ···················· | |
| O | 1305 hCoV-19/England/MILK-A7A623/2020\|EPI_ISL_606627\|20 | ···················· | ···················· | ···················· | ···················· | ···················· | ···················· | |
| O | 1306 hCoV-19/England/MILK-A7AE67/2020\|EPI_ISL_606629\|20 | ···················· | ·A·················· | ···················· | ···················· | ···················· | ···················· | |
| O | 1307 hCoV-19/England/MILK-A7A4B0/2020\|EPI_ISL_606630\|20 | ···················· | ·A·················· | ···················· | ···················· | ···················· | ···A················ | |
| O | 1308 hCoV-19/England/MILK-A7A3E0/2020\|EPI_ISL_606631\|20 | ···················· | ···················· | ···················· | ···················· | ···················· | ···················· | |
| O | 1309 hCoV-19/England/MILK-A7986A/2020\|EPI_ISL_606632\|20 | ···················· | ···················· | ···················· | ···················· | ······T············· | ···················· | |
| O | 1310 hCoV-19/England/MILK-A79581/2020\|EPI_ISL_606635\|20 | ···················· | ···················· | ···················· | ···················· | ···················· | ···················· | |
| O | 1311 hCoV-19/England/MILK-A7D09B/2020\|EPI_ISL_606636\|20 | ···················· | ···················· | ···················· | ···················· | ···················· | ···················· | |
| O | 1312 hCoV-19/England/MILK-A79A28/2020\|EPI_ISL_606637\|20 | ···················· | ···················· | ···················· | ···················· | ···················· | ···················· | |
| O | 1313 hCoV-19/England/MILK-A7ADF1/2020\|EPI_ISL_606638\|20 | ···················· | ·A·················· | ···················· | ···················· | ···················· | ···················· | |
| O | 1314 hCoV-19/England/MILK-A7CD1D/2020\|EPI_ISL_606639\|20 | ···················· | ···················· | ···················· | ···················· | ······T············· | ···················· | |
| O | 1315 hCoV-19/England/MILK-A7AC30/2020\|EPI_ISL_606640\|20 | ···················· | ···················· | ···················· | ···················· | ···················· | ···················· | |
| O | 1316 hCoV-19/England/MILK-A79D2F/2020\|EPI_ISL_606641\|20 | ···················· | ···················· | ···················· | ······G············· | ···················· | ···················· | |
| O | 1317 hCoV-19/England/MILK-A79DD4/2020\|EPI_ISL_606642\|20 | ···················· | ···················· | ···················· | ···················· | ···················· | ···················· | |
| O | 1318 hCoV-19/England/MILK-A7A3C2/2020\|EPI_ISL_606643\|20 | ···················· | ···················· | ···················· | ···················· | ···················· | ···················· | |
| O | 1319 hCoV-19/England/MILK-A7CD2C/2020\|EPI_ISL_606644\|20 | ···················· | ·A·················· | ···················· | ···················· | ···················· | ···A················ | |
| O | 1320 hCoV-19/England/MILK-A79E77/2020\|EPI_ISL_606645\|20 | ···················· | ···················· | ···················· | ···················· | ···················· | ···················· | |
| O | 1321 hCoV-19/England/MILK-A79D7A/2020\|EPI_ISL_606647\|20 | ···················· | ···················· | ···················· | ···················· | ······T············· | ···················· | |
| O | 1322 hCoV-19/England/MILK-A7AC4F/2020\|EPI_ISL_606648\|20 | ···················· | ···················· | ···················· | ···················· | ···················· | ···················· | |
| O | 1323 hCoV-19/England/MILK-A7D56F/2020\|EPI_ISL_606649\|20 | ···················· | ···················· | ···················· | ···················· | ······T············· | ···················· | |
| O | 1324 hCoV-19/England/MILK-A7A2D4/2020\|EPI_ISL_606650\|20 | ···················· | ···················· | ···················· | ···················· | ···················· | ···················· | |
| O | 1325 hCoV-19/England/MILK-A7AC6D/2020\|EPI_ISL_606651\|20 | ···················· | ···················· | ···················· | ···················· | ···················· | ···················· | |
| O | 1326 hCoV-19/England/MILK-A79FCF/2020\|EPI_ISL_606652\|20 | ···················· | ···················· | ···················· | ···················· | ···················· | ···················· | |
| O | 1327 hCoV-19/England/MILK-A7A492/2020\|EPI_ISL_606653\|20 | ···················· | ·A·······A·········· | ···················· | ···················· | ···················· | ···················· | |
| O | 1328 hCoV-19/England/MILK-A79C5F/2020\|EPI_ISL_606656\|20 | ···················· | ···················· | ···················· | ···················· | ···················· | ···················· | |
| O | 1329 hCoV-19/England/MILK-A7A8E1/2020\|EPI_ISL_606657\|20 | ···················· | ···················· | ···················· | ···················· | ···················· | ···················· | |
| O | 1330 hCoV-19/England/MILK-A79B52/2020\|EPI_ISL_606658\|20 | ···················· | ···················· | ···················· | ···················· | ···················· | ···················· | |
| O | 1331 hCoV-19/England/MILK-A7CCC5/2020\|EPI_ISL_606659\|20 | ···················· | ···················· | ···················· | ···················· | ···················· | ···················· | |
| O | 1332 hCoV-19/England/MILK-A7CF80/2020\|EPI_ISL_606660\|20 | ···················· | ···················· | ···················· | ···················· | ···················· | ···················· | |
| O | 1333 hCoV-19/England/MILK-A7985B/2020\|EPI_ISL_606661\|20 | ···················· | ···················· | ···················· | ···················· | ······T············· | ···················· | |
| O | 1334 hCoV-19/England/MILK-A79EB3/2020\|EPI_ISL_606662\|20 | ···················· | ···················· | ···················· | ···················· | ···················· | ···················· | |
| O | 1335 hCoV-19/England/MILK-A79E95/2020\|EPI_ISL_606663\|20 | ···················· | ···················· | ···················· | ···················· | ···················· | ···················· | |
| O | 1336 hCoV-19/England/MILK-A7A8C3/2020\|EPI_ISL_606664\|20 | ···················· | ···················· | ···················· | ···················· | ···················· | ···················· | |
| O | 1337 hCoV-19/England/MILK-A7AB60/2020\|EPI_ISL_606665\|20 | ···················· | ···················· | ···················· | ···················· | ···················· | ···················· | |
| O | 1338 hCoV-19/England/MILK-A7A3FF/2020\|EPI_ISL_606666\|20 | ···················· | ···················· | ···················· | ···················· | ···················· | ···················· | |
| O | 1339 hCoV-19/England/MILK-A79AA0/2020\|EPI_ISL_606667\|20 | ···················· | ···················· | ···················· | ···················· | ···················· | ···················· | |

O 1340 hCoV-19/England/MILK-A7CE1A/2020|EPI_ISL_606668|20 ···················· ···················· ···················· ···················· ···················· ····················
O 1341 hCoV-19/England/MILK-A7978B/2020|EPI_ISL_606669|20 ···················· ···················· ···················· ···················· ···················· ····················
O 1342 hCoV-19/England/MILK-A7CE65/2020|EPI_ISL_606670|20 ···················· ···················· ···················· ···················· ···················· ····················
O 1343 hCoV-19/England/MILK-A79FB0/2020|EPI_ISL_606672|20 ···················· ···················· ···················· ···················· ···················· ····················
O 1344 hCoV-19/England/MILK-A7A3A4/2020|EPI_ISL_606673|20 ···················· ···················· ···················· ···················· ···················· ····················
O 1345 hCoV-19/England/MILK-A7AD00/2020|EPI_ISL_606674|20 ···················· ·A·················· ···················· ···················· ···················· ···A················
O 1346 hCoV-19/England/MILK-A798E2/2020|EPI_ISL_606675|20 ···················· ·R·················· ···················· ···················· ···················· ···A················
O 1347 hCoV-19/England/MILK-A7A104/2020|EPI_ISL_606676|20 ···················· ···················· ···················· ···················· ···················· ····················
O 1348 hCoV-19/England/MILK-A7ABBB/2020|EPI_ISL_606677|20 ···················· ···················· ···················· ···················· ···················· ····················
O 1349 hCoV-19/England/MILK-A7AC9A/2020|EPI_ISL_606678|20 ···················· ···················· ···················· ···················· ···················· ····················
O 1350 hCoV-19/England/MILK-A7CE29/2020|EPI_ISL_606679|20 ···················· ···················· ···················· ···················· ···················· ····················
O 1351 hCoV-19/England/MILK-A7A632/2020|EPI_ISL_606680|20 ···················· ···················· ···················· ···················· ···················· ····················
O 1352 hCoV-19/England/MILK-A7A17D/2020|EPI_ISL_606682|20 ···················· ···················· ···················· ···················· ···················· ····················
O 1353 hCoV-19/England/MILK-A79F38/2020|EPI_ISL_606683|20 ···················· ···················· ···················· ···················· ···················· ····················
O 1354 hCoV-19/England/MILK-A799FE/2020|EPI_ISL_606684|20 ···················· ···················· ···················· ···················· ···················· ····················
O 1355 hCoV-19/England/MILK-A7991C/2020|EPI_ISL_606685|20 ···················· ···················· ···················· ···················· ···················· ····················
O 1356 hCoV-19/England/MILK-A799B2/2020|EPI_ISL_606686|20 ···················· ···················· ···················· ···················· ···················· ····················
O 1357 hCoV-19/England/MILK-A7D07D/2020|EPI_ISL_606687|20 ···················· ···················· ···················· ···················· ···················· ····················
O 1358 hCoV-19/England/MILK-A79ABF/2020|EPI_ISL_606688|20 ···················· ···················· ···················· ···················· ···················· ····················
O 1359 hCoV-19/England/MILK-A7CFAE/2020|EPI_ISL_606689|20 ···················· ···················· ···················· ···················· ···················· ····················
O 1360 hCoV-19/England/MILK-A7A447/2020|EPI_ISL_606690|20 ···················· ···················· ···················· ···················· ···················· ····················
O 1361 hCoV-19/England/MILK-A7A562/2020|EPI_ISL_606691|20 ···················· ···················· ···················· ···················· ······T············· ····················
O 1362 hCoV-19/England/MILK-A79E86/2020|EPI_ISL_606693|20 ···················· ···················· ···················· ···················· ···················· ····················
O 1363 hCoV-19/England/MILK-A79B8F/2020|EPI_ISL_606695|20 ···················· ···················· ···················· ···················· ···················· ····················
O 1364 hCoV-19/England/MILK-A7A508/2020|EPI_ISL_606696|20 ···················· ···················· ···················· ···················· ······T············· ····················
O 1365 hCoV-19/England/MILK-A7A16E/2020|EPI_ISL_606697|20 ···················· ·A·················· ···················· ···················· ···················· ···A················
O 1366 hCoV-19/England/MILK-A79ED1/2020|EPI_ISL_606698|20 ···················· ···················· ···················· ···················· ···················· ····················
O 1367 hCoV-19/England/MILK-A79CC8/2020|EPI_ISL_606699|20 ···················· ···················· ···················· ···················· ······T············· ····················
O 1368 hCoV-19/England/ALDP-A6DFEC/2020|EPI_ISL_607900|20 ···················· ···················· ···················· ···················· ···················· ····················
O 1369 hCoV-19/England/ALDP-A6FA07/2020|EPI_ISL_607901|20 ···················· ···················· ···················· ···················· ···················· ····················
O 1370 hCoV-19/England/ALDP-A6DB9D/2020|EPI_ISL_607902|20 ···················· ···················· ···················· ···················· ···················· ····················
O 1371 hCoV-19/England/ALDP-A6E024/2020|EPI_ISL_607903|20 ···················· ···················· ···················· ···················· ···················· ····················
O 1372 hCoV-19/England/ALDP-A6E200/2020|EPI_ISL_607904|20 ···················· ···················· ···················· ···················· ···················· ····················
O 1373 hCoV-19/England/ALDP-A6F9BF/2020|EPI_ISL_607905|20 ···················· ···················· ···················· ···················· ···················· ····················
O 1374 hCoV-19/England/ALDP-A703EA/2020|EPI_ISL_607950|20 ···················· ···················· ···················· ···················· ···················· ····················
O 1375 hCoV-19/England/ALDP-A6E9CF/2020|EPI_ISL_607951|20 ···················· ···················· ···················· ···················· ···················· ····················
O 1376 hCoV-19/England/ALDP-A6DADC/2020|EPI_ISL_607952|20 ···················· ···················· ···················· ···················· ···················· ····················
O 1377 hCoV-19/England/ALDP-A6E73E/2020|EPI_ISL_607953|20 ···················· ···················· ···················· ···················· ···················· ····················
O 1378 hCoV-19/England/ALDP-A6EA62/2020|EPI_ISL_607954|20 ···················· ···················· ···················· ···················· ···················· ····················
O 1379 hCoV-19/England/ALDP-A6F58E/2020|EPI_ISL_607955|20 ···················· ···················· ···················· ···················· ···················· ····················
O 1380 hCoV-19/England/ALDP-A6F57F/2020|EPI_ISL_607956|20 ···················· ···················· ···················· ···················· ···················· ····················
O 1381 hCoV-19/England/ALDP-A6DB15/2020|EPI_ISL_607957|20 ···················· ···················· ···················· ···················· ···················· ····················
O 1382 hCoV-19/England/ALDP-A6F3C0/2020|EPI_ISL_607958|20 ···················· ···················· ···················· ···················· ···················· ····················
O 1383 hCoV-19/England/ALDP-A6DFFB/2020|EPI_ISL_607959|20 ···················· ···················· ···················· ···················· ···················· ····················
O 1384 hCoV-19/England/ALDP-A6E437/2020|EPI_ISL_607961|20 ···················· ···················· ···················· ···················· ···················· ····················
O 1385 hCoV-19/England/ALDP-A6F409/2020|EPI_ISL_607962|20 ···················· ···················· ···················· ···················· ···················· ····················
O 1386 hCoV-19/England/ALDP-A6F17B/2020|EPI_ISL_607963|20 ···················· ···················· ···················· ···················· ···················· ····················
O 1387 hCoV-19/England/ALDP-A6E938/2020|EPI_ISL_607964|20 ···················· ···················· ···················· ···················· ···················· ····················
O 1388 hCoV-19/England/ALDP-A6E710/2020|EPI_ISL_607965|20 ···················· ···················· ···················· ···················· ···················· ····················
O 1389 hCoV-19/England/ALDP-A6F032/2020|EPI_ISL_607967|20 ···················· ···················· ···················· ···················· ···················· ····················
O 1390 hCoV-19/England/ALDP-A6EA08/2020|EPI_ISL_607968|20 ···················· ···················· ···················· ···················· ···················· ····················
O 1391 hCoV-19/England/ALDP-A6EA71/2020|EPI_ISL_607969|20 ···················· ···················· ···················· ···················· ···················· ····················
O 1392 hCoV-19/England/ALDP-A6E868/2020|EPI_ISL_607970|20 ···················· ···················· ···················· ···················· ···················· ····················
O 1393 hCoV-19/England/ALDP-A6DB8E/2020|EPI_ISL_607971|20 ···················· ···················· ···················· ···················· ···················· ····················
O 1394 hCoV-19/England/ALDP-A6F6A9/2020|EPI_ISL_607972|20 ···················· ···················· ···················· ···················· ···················· ····················
O 1395 hCoV-19/England/ALDP-A6E9ED/2020|EPI_ISL_607973|20 ···················· ·········T·········· ···················· ···················· ···················· ····················
O 1396 hCoV-19/England/ALDP-A704AB/2020|EPI_ISL_607974|20 ···················· ···················· ···················· ···················· ···················· ····················
O 1397 hCoV-19/England/ALDP-A70432/2020|EPI_ISL_607975|20 ···················· ···················· ···················· ···················· ···················· ····················
O 1398 hCoV-19/England/ALDP-A6EF27/2020|EPI_ISL_607976|20 ···················· ···················· ···················· ···················· ···················· ····················
O 1399 hCoV-19/England/ALDP-A6FA25/2020|EPI_ISL_607977|20 ···················· ···················· ···················· ···················· ···················· ····················
O 1400 hCoV-19/England/ALDP-A6F4DC/2020|EPI_ISL_607980|20 ···················· ···················· ···················· ···················· ···················· ····················
O 1401 hCoV-19/England/ALDP-A7072A/2020|EPI_ISL_607981|20 ···················· ···················· ···················· ···················· ···················· ····················
O 1402 hCoV-19/England/ALDP-A705D5/2020|EPI_ISL_607982|20 ···················· ···················· ···················· ···················· ···················· ····················
O 1403 hCoV-19/England/ALDP-A6C32D/2020|EPI_ISL_607983|20 ···················· ···················· ···················· ···················· ···················· ····················
O 1404 hCoV-19/England/ALDP-A6D1AA/2020|EPI_ISL_607984|20 ···················· ···················· ···················· ···················· ···················· ····················
O 1405 hCoV-19/England/ALDP-A6D483/2020|EPI_ISL_607985|20 ···················· ···················· ···················· ···················· ···················· ····················
O 1406 hCoV-19/England/ALDP-A6C8B5/2020|EPI_ISL_607986|20 ···················· ···················· ···················· ···················· ···················· ····················
O 1407 hCoV-19/England/ALDP-A6D0AD/2020|EPI_ISL_607987|20 ···················· ···················· ···················· ···················· ···················· ····················
O 1408 hCoV-19/England/ALDP-A70238/2020|EPI_ISL_607988|20 ···················· ···················· ···················· ···················· ···················· ····················
O 1409 hCoV-19/England/ALDP-A6C6E8/2020|EPI_ISL_607989|20 ···················· ···················· ···················· ···················· ···················· ····················
O 1410 hCoV-19/England/ALDP-A6C83D/2020|EPI_ISL_607990|20 ···················· ···················· ···················· ···················· ···················· ····················
O 1411 hCoV-19/Wales/ALDP-A6CE68/2020|EPI_ISL_607991|2020 ···················· ···················· ···················· ···················· ···················· ····················
O 1412 hCoV-19/England/ALDP-A6CDA7/2020|EPI_ISL_607992|20 ···················· ···················· ···················· ···················· ···················· ····················
O 1413 hCoV-19/England/ALDP-A7005C/2020|EPI_ISL_607993|20 ···················· ···················· ···················· ···················· ···················· ····················
O 1414 hCoV-19/England/ALDP-A6C581/2020|EPI_ISL_607995|20 ···················· ···················· ···················· ···················· ···················· ····················
O 1415 hCoV-19/England/ALDP-A6C527/2020|EPI_ISL_607997|20 ···················· ···················· ···················· ···················· ···················· ····················
O 1416 hCoV-19/England/ALDP-A6CD5C/2020|EPI_ISL_607998|20 ···················· ·A·················· ···················· ···················· ···················· ····················
O 1417 hCoV-19/England/ALDP-A6D429/2020|EPI_ISL_607999|20 ···················· ···················· ···················· ···················· ···················· ····················
O 1418 hCoV-19/England/ALDP-A923B5/2020|EPI_ISL_611100|20 ···················· ···················· ···················· ···················· ···················· ····················
O 1419 hCoV-19/England/ALDP-A92D5D/2020|EPI_ISL_611101|20 ···················· ···················· ···················· ···················· ···················· ····················

| | | | | | | | | | |
|---|---|---|---|---|---|---|---|---|---|
| O | 1420 | hCoV-19/England/ALDP-A9368E/2020\|EPI_ISL_611102\|20 | ···················· | ···················· | ···················· | ···················· | ···················· | ···················· | |
| O | 1421 | hCoV-19/England/ALDP-A93967/2020\|EPI_ISL_611104\|20 | ···················· | ···················· | ···················· | ···················· | ···················· | ···················· | |
| O | 1422 | hCoV-19/England/ALDP-A93A46/2020\|EPI_ISL_611105\|20 | ···················· | ···················· | ···················· | ···················· | ···················· | ···················· | |
| O | 1423 | hCoV-19/England/ALDP-A92E3C/2020\|EPI_ISL_611106\|20 | ···················· | ···················· | ···················· | ···················· | ···················· | ···················· | |
| O | 1424 | hCoV-19/England/ALDP-A93DE3/2020\|EPI_ISL_611107\|20 | ···················· | ···················· | ···················· | ···················· | ···················· | ···················· | |
| O | 1425 | hCoV-19/England/ALDP-A927AA/2020\|EPI_ISL_611108\|20 | ···················· | ···················· | ···················· | ···················· | ···················· | ···················· | |
| O | 1426 | hCoV-19/England/ALDP-A9440B/2020\|EPI_ISL_611109\|20 | ···················· | ·A·················· | ···················· | ···················· | ···················· | ···················· | |
| O | 1427 | hCoV-19/England/ALDP-A948C3/2020\|EPI_ISL_611110\|20 | ···················· | ···················· | ···················· | ···················· | ···················· | ···················· | |
| O | 1428 | hCoV-19/England/ALDP-A928B6/2020\|EPI_ISL_611112\|20 | ···················· | ···················· | ···················· | ···················· | ···················· | ···················· | |
| O | 1429 | hCoV-19/England/ALDP-A939C1/2020\|EPI_ISL_611113\|20 | ···················· | ···················· | ···················· | ···················· | ···················· | ···················· | |
| O | 1430 | hCoV-19/England/ALDP-A93ACE/2020\|EPI_ISL_611114\|20 | ···················· | ···················· | ···················· | ···················· | ···················· | ···················· | |
| O | 1431 | hCoV-19/England/ALDP-A927B9/2020\|EPI_ISL_611115\|20 | ···················· | ···················· | ···················· | ···················· | ···················· | ···················· | |
| O | 1432 | hCoV-19/England/ALDP-A93B52/2020\|EPI_ISL_611116\|20 | ···················· | ···················· | ···················· | ···················· | ···················· | ···················· | |
| O | 1433 | hCoV-19/England/ALDP-A92D7B/2020\|EPI_ISL_611119\|20 | ···················· | ···················· | ···················· | ···················· | ···················· | ···················· | |
| O | 1434 | hCoV-19/England/ALDP-A93B70/2020\|EPI_ISL_611120\|20 | ···················· | ···················· | ···················· | ···················· | ···················· | ···················· | |
| O | 1435 | hCoV-19/England/ALDP-A92E69/2020\|EPI_ISL_611121\|20 | ···················· | ···················· | ···················· | ···················· | ···················· | ···················· | |
| O | 1436 | hCoV-19/England/ALDP-A93C9B/2020\|EPI_ISL_611122\|20 | ···················· | ···················· | ···················· | ···················· | ···················· | ···················· | |
| O | 1437 | hCoV-19/England/ALDP-A92D6C/2020\|EPI_ISL_611123\|20 | ···················· | ···················· | ···················· | ···················· | ···················· | ···················· | |
| O | 1438 | hCoV-19/England/ALDP-A93F65/2020\|EPI_ISL_611124\|20 | ···················· | ···················· | ···················· | ···················· | ···················· | ···················· | |
| O | 1439 | hCoV-19/England/ALDP-A926DA/2020\|EPI_ISL_611125\|20 | ···················· | ···················· | ···················· | ···················· | ···················· | ···················· | |
| O | 1440 | hCoV-19/England/ALDP-A93C04/2020\|EPI_ISL_611127\|20 | ···················· | ···················· | ···················· | ···················· | ···················· | ···················· | |
| O | 1441 | hCoV-19/England/ALDP-A93F47/2020\|EPI_ISL_611128\|20 | ···················· | ···················· | ···················· | ···················· | ···················· | ···················· | |
| O | 1442 | hCoV-19/England/ALDP-A928C5/2020\|EPI_ISL_611140\|20 | ···················· | ···················· | ···················· | ···················· | ···················· | ···················· | |
| O | 1443 | hCoV-19/England/ALDP-A93FFC/2020\|EPI_ISL_611142\|20 | ···················· | ···················· | ···················· | ···················· | ···················· | ···················· | |
| O | 1444 | hCoV-19/England/ALDP-A92CE7/2020\|EPI_ISL_611147\|20 | ···················· | ···················· | ···················· | ···················· | ···················· | ···················· | |
| O | 1445 | hCoV-19/England/ALDP-A92D4E/2020\|EPI_ISL_611149\|20 | ···················· | ···················· | ···················· | ···················· | ···················· | ···················· | |
| O | 1446 | hCoV-19/England/ALDP-A92B17/2020\|EPI_ISL_611150\|20 | ···················· | ···················· | ···················· | ···················· | ···················· | ···················· | |
| O | 1447 | hCoV-19/England/ALDP-A92D11/2020\|EPI_ISL_611151\|20 | ···················· | ···················· | ···················· | ···················· | ···················· | ···················· | |
| O | 1448 | hCoV-19/England/ALDP-A943B3/2020\|EPI_ISL_611154\|20 | ···················· | ···················· | ···················· | ···················· | ···················· | ···················· | |
| O | 1449 | hCoV-19/England/ALDP-A96E92/2020\|EPI_ISL_611155\|20 | ···················· | ···················· | ···················· | ···················· | ···················· | ···················· | |
| O | 1450 | hCoV-19/England/ALDP-A924FE/2020\|EPI_ISL_611157\|20 | ···················· | ···················· | ···················· | ···················· | ···················· | ···················· | |
| O | 1451 | hCoV-19/England/ALDP-A93D3E/2020\|EPI_ISL_611158\|20 | ···················· | ···················· | ···················· | ···················· | ···················· | ···················· | |
| O | 1452 | hCoV-19/England/ALDP-A93F0B/2020\|EPI_ISL_611159\|20 | ···················· | ···················· | ···················· | ···················· | ···················· | ···················· | |
| O | 1453 | hCoV-19/England/ALDP-A923E2/2020\|EPI_ISL_611160\|20 | ···················· | ···················· | ···················· | ···················· | ···················· | ···················· | |
| O | 1454 | hCoV-19/England/ALDP-A93C22/2020\|EPI_ISL_611161\|20 | ···················· | ···················· | ···················· | ···················· | ···················· | ···················· | |
| O | 1455 | hCoV-19/England/ALDP-A9282F/2020\|EPI_ISL_611162\|20 | ···················· | ···················· | ···················· | ···················· | ···················· | ···················· | |
| O | 1456 | hCoV-19/England/ALDP-A93642/2020\|EPI_ISL_611163\|20 | ···················· | ···················· | ···················· | ···················· | ···················· | ···················· | |
| O | 1457 | hCoV-19/England/ALDP-A926BC/2020\|EPI_ISL_611164\|20 | ···················· | ···················· | ···················· | ···················· | ···················· | ···················· | |
| O | 1458 | hCoV-19/England/ALDP-A93E86/2020\|EPI_ISL_611165\|20 | ···················· | ···················· | ···················· | ···················· | ···················· | ···················· | |
| O | 1459 | hCoV-19/England/ALDP-A9485A/2020\|EPI_ISL_611166\|20 | ···················· | ···················· | ···················· | ···················· | ···················· | ···················· | |
| O | 1460 | hCoV-19/England/ALDP-A92DA8/2020\|EPI_ISL_611167\|20 | ···················· | ···················· | ···················· | ···················· | ···················· | ···················· | |
| O | 1461 | hCoV-19/England/ALDP-A92731/2020\|EPI_ISL_611169\|20 | ···················· | ···················· | ···················· | ···················· | ···················· | ···················· | |
| O | 1462 | hCoV-19/England/LIVE-1D2CD5/2020\|EPI_ISL_612416\|20 | ···················· | ···················· | ···················· | ···················· | ···················· | ···················· | |
| O | 1463 | hCoV-19/England/LIVE-1D2D1E/2020\|EPI_ISL_612417\|20 | ···················· | ···················· | ···················· | ···················· | ···················· | ···················· | |
| O | 1464 | hCoV-19/England/LIVE-1D2D2D/2020\|EPI_ISL_612418\|20 | ···················· | ···················· | ···················· | ···················· | ···················· | ···················· | |
| O | 1465 | hCoV-19/England/LIVE-1D2D4B/2020\|EPI_ISL_612419\|20 | ···················· | ···················· | ···················· | ···················· | ···················· | ···················· | |
| O | 1466 | hCoV-19/England/LIVE-1D2D5A/2020\|EPI_ISL_612420\|20 | ···················· | ···················· | ···················· | ···················· | ···················· | ···················· | |
| O | 1467 | hCoV-19/England/LIVE-1D2D69/2020\|EPI_ISL_612421\|20 | ···················· | ···················· | ···················· | ···················· | ···················· | ···················· | |
| O | 1468 | hCoV-19/England/LIVE-1D2DA5/2020\|EPI_ISL_612422\|20 | ···················· | ···················· | ···················· | ···················· | ···················· | ···················· | |
| O | 1469 | hCoV-19/England/LIVE-1D2DD2/2020\|EPI_ISL_612423\|20 | ···················· | ···················A | ···················· | ···················· | ···················· | ···················· | 1 |
| O | 1470 | hCoV-19/England/LIVE-1D2DE1/2020\|EPI_ISL_612424\|20 | ···················· | ···················· | ···················· | ···················· | ···················· | ···················· | |
| O | 1471 | hCoV-19/England/LIVE-1D2DF0/2020\|EPI_ISL_612425\|20 | ···················· | ···················· | ···················· | ···················· | ···················· | ···················· | |
| O | 1472 | hCoV-19/England/LIVE-1D2E0C/2020\|EPI_ISL_612426\|20 | ···················· | ···················· | ···················· | ···················· | ···················· | ···················· | |
| O | 1473 | hCoV-19/England/LIVE-1D2E2A/2020\|EPI_ISL_612427\|20 | ···················· | ···················· | ···················· | ···················· | ···················· | ···················· | |
| O | 1474 | hCoV-19/England/LIVE-1D2E39/2020\|EPI_ISL_612428\|20 | ···················· | ···················· | ···················· | ···················· | ···················· | ···················· | |
| O | 1475 | hCoV-19/England/LIVE-1D2E48/2020\|EPI_ISL_612429\|20 | ···················· | ···················· | ···················· | ···················· | ···················· | ···················· | |
| O | 1476 | hCoV-19/England/LIVE-1D2E57/2020\|EPI_ISL_612430\|20 | ···················· | ···················· | ···················· | ···················· | ···················· | ···················· | |
| O | 1477 | hCoV-19/England/LIVE-1D2E93/2020\|EPI_ISL_612431\|20 | ···················· | ···················· | ···················· | ···················· | ···················· | ···················· | |
| O | 1478 | hCoV-19/England/LIVE-1D2EB1/2020\|EPI_ISL_612432\|20 | ···················· | ···················· | ···················· | ···················· | ···················· | ···················· | |
| O | 1479 | hCoV-19/England/LIVE-1DD627/2020\|EPI_ISL_612433\|20 | ···················· | ···················· | ···················· | ···················· | ···················· | ···················· | |
| O | 1480 | hCoV-19/England/LIVE-1DD7AC/2020\|EPI_ISL_612434\|20 | ···················· | ···················· | ···················· | ···················· | ···················· | ···················· | |
| O | 1481 | hCoV-19/England/LIVE-1DD821/2020\|EPI_ISL_612435\|20 | ···················· | ···················· | ···················· | ···················· | ···················· | ···················· | |
| O | 1482 | hCoV-19/England/LIVE-1DD95B/2020\|EPI_ISL_612436\|20 | ···················· | ···················· | ···················· | ···················· | ···················· | ···················· | |
| O | 1483 | hCoV-19/England/LIVE-AB571/2020\|EPI_ISL_612437\|202 | ···················· | ···················· | ···················· | ···················· | ···················· | ···················· | |
| O | 1484 | hCoV-19/England/LIVE-AB5CC/2020\|EPI_ISL_612438\|202 | ···················· | ···················· | ···················· | ···················· | ···················· | ···················· | |
| O | 1485 | hCoV-19/England/LIVE-AB69C/2020\|EPI_ISL_612439\|202 | ···················· | ···················· | ···················· | ···················· | ···················· | ···················· | |
| O | 1486 | hCoV-19/England/LIVE-AB76C/2020\|EPI_ISL_612440\|202 | ···················· | ···················· | ···················· | ···················· | ···················· | ···················· | |
| O | 1487 | hCoV-19/England/LIVE-AB896/2020\|EPI_ISL_612441\|202 | ···················· | ···················· | ···················· | ···················· | ···················· | ···················· | |
| O | 1488 | hCoV-19/England/LIVE-AB8B4/2020\|EPI_ISL_612442\|202 | ···················· | ···················· | ···················· | ···················· | ···················· | ···················· | |
| O | 1489 | hCoV-19/England/LIVE-AB948/2020\|EPI_ISL_612443\|202 | ···················· | ···················· | ···················· | ···················· | ···················· | ···················· | |
| O | 1490 | hCoV-19/England/LIVE-AB975/2020\|EPI_ISL_612444\|202 | ···················· | ···················· | ···················· | ···················· | ···················· | ···················· | |
| O | 1491 | hCoV-19/England/LIVE-AB9A2/2020\|EPI_ISL_612445\|202 | ···················· | ···················· | ···················· | ···················· | ···················· | ···················· | |
| O | 1492 | hCoV-19/England/LIVE-AB9C0/2020\|EPI_ISL_612446\|202 | ···················· | ···················· | ···················· | ···················· | ···················· | ···················· | |
| O | 1493 | hCoV-19/England/LIVE-AB9EE/2020\|EPI_ISL_612447\|202 | ···················· | ···················· | ···················· | ···················· | ···················· | ···················· | |
| O | 1494 | hCoV-19/England/LIVE-ABA18/2020\|EPI_ISL_612448\|202 | ···················· | ···················· | ···················· | ···················· | ···················· | ···················· | |
| O | 1495 | hCoV-19/England/LOND-1262E94/2020\|EPI_ISL_612450\|2 | ···················· | ···················· | ···················· | ···················· | ···················· | ···················· | |
| O | 1496 | hCoV-19/England/LOND-1262F0A/2020\|EPI_ISL_612451\|2 | ···················· | ···················· | ···················· | ···················· | ···················· | ···················· | |
| O | 1497 | hCoV-19/England/LOND-D9B69/2020\|EPI_ISL_612452\|202 | ···················· | ···················· | ···················· | ···················· | ···················· | ···················· | |
| O | 1498 | hCoV-19/England/LOND-D9C48/2020\|EPI_ISL_612453\|202 | ···················· | ···················· | ···················· | ···················· | ···················· | ···················· | |
| O | 1499 | hCoV-19/England/LOND-D9C57/2020\|EPI_ISL_612454\|202 | ···················· | ···················· | ···················· | ···················· | ···················· | ···················· | |

| | | | | | | | | |
|---|---|---|---|---|---|---|---|---|
| O | 1500 hCoV-19/England/LOND-D9C66/2020\|EPI_ISL_612455\|202 | ···················· | ·A·················· | ···················· | ···················· | ···················· | ···················· | |
| O | 1501 hCoV-19/England/LOND-D9CEE/2020\|EPI_ISL_612456\|202 | ···················· | ···················· | ···················· | ···················· | ·····T·············· | ···················· | |
| O | 1502 hCoV-19/England/MILK-A1885E/2020\|EPI_ISL_612457\|20 | ···················· | ···················· | ···················· | ···················· | ···················· | ···················· | |
| O | 1503 hCoV-19/England/MILK-A18D5F/2020\|EPI_ISL_612458\|20 | ···················· | ···················· | ···················· | ···················· | ·····T·············· | ···················· | |
| O | 1504 hCoV-19/England/MILK-A18DB9/2020\|EPI_ISL_612459\|20 | ···················· | ···················· | ···················· | ···················· | ···················· | ···················· | |
| O | 1505 hCoV-19/England/MILK-A18E7A/2020\|EPI_ISL_612470\|20 | ···················· | ···················· | ···················· | ···················· | ·····T·············· | ···················· | |
| O | 1506 hCoV-19/England/MILK-A18EA7/2020\|EPI_ISL_612471\|20 | ···················· | ···················· | ···················· | ···················· | ···················· | ···················· | |
| O | 1507 hCoV-19/England/MILK-A18ED4/2020\|EPI_ISL_612472\|20 | ···················· | ···················· | ···················· | ···················· | ···················· | ···················· | |
| O | 1508 hCoV-19/England/MILK-A18EE3/2020\|EPI_ISL_612473\|20 | ···················· | ···················· | ···················· | ···················· | ···················· | ···················· | |
| O | 1509 hCoV-19/England/MILK-A19082/2020\|EPI_ISL_612478\|20 | ···················· | ···················· | ···················· | ···················· | ···················· | ···················· | |
| O | 1510 hCoV-19/England/MILK-A19091/2020\|EPI_ISL_612479\|20 | ···················· | ···················· | ···················· | ···················· | ···················· | ···················· | |
| O | 1512 hCoV-19/England/MILK-A190CE/2020\|EPI_ISL_612481\|20 | ···················· | ···················· | ···················· | ···················· | ···················· | ···················· | |
| O | 1513 hCoV-19/England/MILK-A190EC/2020\|EPI_ISL_612482\|20 | ···················· | ·A·················· | ···················· | ···················· | ···················· | ···················· | |
| O | 1514 hCoV-19/England/MILK-A190FB/2020\|EPI_ISL_612483\|20 | ···················· | ···················· | ···················· | ···················· | ···················· | ···················· | |
| O | 1515 hCoV-19/England/MILK-A19107/2020\|EPI_ISL_612484\|20 | ···················· | ···················· | ···················· | ···················· | ···················· | ···················· | |
| O | 1517 hCoV-19/England/MILK-A19152/2020\|EPI_ISL_612486\|20 | ···················· | ···················· | ···················· | ···················· | ···················· | ···················· | |
| O | 1518 hCoV-19/England/MILK-A1919E/2020\|EPI_ISL_612487\|20 | ···················· | ·R·················· | ···················· | ···················· | ···················· | ··R················· | |
| O | 1519 hCoV-19/England/MILK-A191BC/2020\|EPI_ISL_612488\|20 | ···················· | ···················· | ···················· | ···················· | ···················· | ···················· | |
| O | 1520 hCoV-19/England/MILK-A191CB/2020\|EPI_ISL_612489\|20 | ···················· | ···················· | ···················· | ···················· | ···················· | ···················· | |
| O | 1521 hCoV-19/England/MILK-A191DA/2020\|EPI_ISL_612490\|20 | ···················· | ···················· | ···················· | ···················· | ···················· | ···················· | |
| O | 1522 hCoV-19/England/MILK-A191F8/2020\|EPI_ISL_612491\|20 | ···················· | ···················· | ···················· | ···················· | ···················· | ···················· | |
| O | 1523 hCoV-19/England/MILK-A19204/2020\|EPI_ISL_612492\|20 | ···················· | ·A·················· | ···················· | ···················· | ···················· | ··A················· | |
| O | 1524 hCoV-19/England/MILK-A19231/2020\|EPI_ISL_612493\|20 | ···················· | ·A·················· | ···················· | ···················· | ···················· | ···················· | |
| O | 1525 hCoV-19/England/MILK-A19240/2020\|EPI_ISL_612494\|20 | ···················· | ·A·················· | ···················· | ···················· | ···················· | ··A················· | |
| O | 1526 hCoV-19/England/MILK-A1928C/2020\|EPI_ISL_612495\|20 | ···················· | ···················· | ···················· | ···················· | ···················· | ···················· | |
| O | 1527 hCoV-19/England/MILK-A192AA/2020\|EPI_ISL_612496\|20 | ···················· | ···················· | ···················· | ···················· | ···················· | ···················· | |
| O | 1528 hCoV-19/England/MILK-A192B9/2020\|EPI_ISL_612497\|20 | ···················· | ···················· | ···················· | ···················· | ···················· | ···················· | |
| O | 1530 hCoV-19/England/MILK-A192D7/2020\|EPI_ISL_612499\|20 | ···················· | ·A·················· | ···················· | ···················· | ···················· | ··A················· | |
| O | 1531 hCoV-19/Gibraltar/204421241/2020\|EPI_ISL_613700\|20 | ···················· | ···················· | ···················· | ···················· | ···················· | ···················· | |
| O | 1532 hCoV-19/Gibraltar/204421242/2020\|EPI_ISL_613701\|20 | ···················· | ···················· | ···················· | ···················· | ···················· | ···················· | |
| O | 1533 hCoV-19/Gibraltar/204421243/2020\|EPI_ISL_613702\|20 | ···················· | ···················· | ···················· | ···················· | ···················· | ···················· | |
| O | 1535 hCoV-19/USA/FL-BPHL-1896/2020\|EPI_ISL_613770\|2020- | ···················· | ···················· | ···················· | ···················· | ···················· | ···················· | |
| O | 1536 hCoV-19/USA/FL-BPHL-1915/2020\|EPI_ISL_613771\|2020- | ···················· | ···················· | ···················· | ···················· | ···················· | ···················· | |
| O | 1537 hCoV-19/England/ALDP-A6D2E3/2020\|EPI_ISL_608200\|20 | ···················· | ···················· | ···················· | ···················· | ···················· | ···················· | |
| O | 1538 hCoV-19/England/ALDP-A6D3D1/2020\|EPI_ISL_608201\|20 | ···················· | ···················· | ···················· | ···················· | ···················· | ···················· | |
| O | 1539 hCoV-19/England/ALDP-A6D19B/2020\|EPI_ISL_608203\|20 | ···················· | ···················· | ···················· | ···················· | ···················· | ···················· | |
| O | 1540 hCoV-19/England/ALDP-A6C554/2020\|EPI_ISL_608204\|20 | ···················· | ···················· | ···················· | ···················· | ···················· | ···················· | |
| O | 1541 hCoV-19/England/ALDP-A708CD/2020\|EPI_ISL_608205\|20 | ···················· | ···················· | ···················· | ···················· | ···················· | ···················· | |
| O | 1542 hCoV-19/England/ALDP-A6D878/2020\|EPI_ISL_608206\|20 | ···················· | ···················· | ···················· | ···················· | ···················· | ···················· | |
| O | 1543 hCoV-19/England/ALDP-A6C4A2/2020\|EPI_ISL_608207\|20 | ···················· | ···················· | ···················· | ···················· | ···················· | ···················· | |
| O | 1544 hCoV-19/England/ALDP-A6D517/2020\|EPI_ISL_608208\|20 | ···················· | ···················· | ···················· | ···················· | ···················· | ···················· | |
| O | 1545 hCoV-19/England/ALDP-A6C378/2020\|EPI_ISL_608209\|20 | ···················· | ···················· | ···················· | ···················· | ···················· | ···················· | |
| O | 1546 hCoV-19/England/ALDP-A70089/2020\|EPI_ISL_608212\|20 | ···················· | ···················· | ···················· | ···················· | ···················· | ···················· | |
| O | 1547 hCoV-19/England/ALDP-A6CDD4/2020\|EPI_ISL_608213\|20 | ···················· | ···················· | ···················· | ···················· | ···················· | ···················· | |
| O | 1548 hCoV-19/England/ALDP-A6D22F/2020\|EPI_ISL_608215\|20 | ···················· | ···················· | ···················· | ···················· | ···················· | ···················· | |
| O | 1549 hCoV-19/England/ALDP-A6C730/2020\|EPI_ISL_608217\|20 | ···················· | ···················· | ···················· | ···················· | ···················· | ···················· | |
| O | 1550 hCoV-19/England/ALDP-A70809/2020\|EPI_ISL_608218\|20 | ···················· | ···················· | ···················· | ···················· | ···················· | ···················· | |
| O | 1551 hCoV-19/England/ALDP-A6C86A/2020\|EPI_ISL_608219\|20 | ···················· | ·A·················· | ···················· | ···················· | ···················· | ··A················· | |
| O | 1552 hCoV-19/England/ALDP-A6D2B6/2020\|EPI_ISL_608220\|20 | ···················· | ···················· | ···················· | ···················· | ···················· | ···················· | |
| O | 1553 hCoV-19/England/ALDP-A6CEC2/2020\|EPI_ISL_608221\|20 | ···················· | ···················· | ···················· | ···················· | ···················· | ···················· | |
| O | 1554 hCoV-19/England/ALDP-A6FEFC/2020\|EPI_ISL_608222\|20 | ···················· | ···················· | ···················· | ···················· | ···················· | ···················· | |
| O | 1555 hCoV-19/England/ALDP-A6D24D/2020\|EPI_ISL_608223\|20 | ···················· | ···················· | ···················· | ···················· | ···················· | ···················· | |
| O | 1556 hCoV-19/England/ALDP-A6CF47/2020\|EPI_ISL_608224\|20 | ···················· | ···················· | ···················· | ···················· | ···················· | ···················· | |
| O | 1557 hCoV-19/England/ALDP-A70265/2020\|EPI_ISL_608225\|20 | ···················· | ···················· | ···················· | ···················· | ···················· | ···················· | |
| O | 1558 hCoV-19/England/ALDP-A6D386/2020\|EPI_ISL_608227\|20 | ···················· | ···················· | ···················· | ···················· | ···················· | ···················· | |
| O | 1559 hCoV-19/England/ALDP-A6CCB9/2020\|EPI_ISL_608228\|20 | ···················· | ···················· | ···················· | ···················· | ···················· | ···················· | |
| O | 1560 hCoV-19/England/ALDP-A6D052/2020\|EPI_ISL_608229\|20 | ···················· | ···················· | ···················· | ···················· | ···················· | ···················· | |
| O | 1561 hCoV-19/England/ALDP-A6D7D5/2020\|EPI_ISL_608230\|20 | ···················· | ···················· | ···················· | ···················· | ···················· | ···················· | |
| O | 1562 hCoV-19/England/ALDP-A70186/2020\|EPI_ISL_608231\|20 | ···················· | ···················· | ···················· | ···················· | ···················· | ···················· | |
| O | 1563 hCoV-19/England/ALDP-A6C633/2020\|EPI_ISL_608232\|20 | ···················· | ···················· | ···················· | ···················· | ···················· | ···················· | |
| O | 1564 hCoV-19/England/ALDP-A6FF53/2020\|EPI_ISL_608233\|20 | ···················· | ···················· | ···················· | ···················· | ···················· | ···················· | |
| O | 1565 hCoV-19/England/ALDP-A708AF/2020\|EPI_ISL_608234\|20 | ···················· | ···················· | ···················· | ···················· | ···················· | ···················· | |
| O | 1566 hCoV-19/England/ALDP-A6CEA4/2020\|EPI_ISL_608235\|20 | ··················T· | ···················· | ···················· | ···················· | ···················· | ···················· | 1 |
| O | 1567 hCoV-19/England/ALDP-A6C518/2020\|EPI_ISL_608236\|20 | ···················· | ···················· | ···················· | ···················· | ···················· | ···················· | |
| O | 1568 hCoV-19/England/ALDP-A6D4B0/2020\|EPI_ISL_608238\|20 | ···················· | ···················· | ···················· | ···················· | ···················· | ···················· | |
| O | 1569 hCoV-19/England/ALDP-A6C5DC/2020\|EPI_ISL_608239\|20 | ···················· | ···················· | ···················· | ···················· | ·····T·············· | ···················· | |
| O | 1570 hCoV-19/England/ALDP-A6CFDE/2020\|EPI_ISL_608284\|20 | ···················· | ···················· | ···················· | ···················· | ···················· | ···················· | |
| O | 1571 hCoV-19/England/ALDP-A6CBAD/2020\|EPI_ISL_608285\|20 | ···················· | ···················· | ···················· | ···················· | ···················· | ···················· | |
| O | 1572 hCoV-19/England/CAMC-A418C5/2020\|EPI_ISL_609500\|20 | ···················· | ·A·················· | ···················· | ···················· | ···················· | ··A················· | |
| O | 1573 hCoV-19/England/CAMC-A41E3C/2020\|EPI_ISL_609501\|20 | ···················· | ···················· | ···················· | ···················· | ···················· | ···················· | |
| O | 1574 hCoV-19/England/CAMC-A420DB/2020\|EPI_ISL_609502\|20 | ···················· | ···················· | ···················· | ···················· | ···················· | ···················· | |
| O | 1575 hCoV-19/England/CAMC-A41B26/2020\|EPI_ISL_609503\|20 | ···················· | ·A·················· | ···················· | ···················· | ···················· | ···················· | |
| O | 1576 hCoV-19/England/CAMC-A40D03/2020\|EPI_ISL_609504\|20 | ···················· | ···················· | ···················· | ···················· | ···················· | ···················· | |
| O | 1577 hCoV-19/England/CAMC-A40FFE/2020\|EPI_ISL_609505\|20 | ···················· | ···················· | ···················· | ···················· | ···················· | ···················· | |
| O | 1578 hCoV-19/England/CAMC-A40EA6/2020\|EPI_ISL_609509\|20 | ···················· | ···················· | ···················· | ···················· | ···················· | ···················· | |
| O | 1579 hCoV-19/England/CAMC-A40EC4/2020\|EPI_ISL_609511\|20 | ···················· | ···················· | ···················· | ···················· | ···················· | ···················· | |
| O | 1580 hCoV-19/England/CAMC-A41EB4/2020\|EPI_ISL_609513\|20 | ···················· | ···················· | ···················· | ···················· | ···················· | ···················· | |
| O | 1581 hCoV-19/England/CAMC-A40D5E/2020\|EPI_ISL_609514\|20 | ···················· | ···················· | ···················· | ···················· | ···················· | ···················· | |
| O | 1582 hCoV-19/England/CAMC-A420CC/2020\|EPI_ISL_609515\|20 | ···················· | ···················· | ···················· | ···················· | ···················· | ···················· | |
| O | 1583 hCoV-19/England/CAMC-A41CBA/2020\|EPI_ISL_609516\|20 | ···················· | ···················· | ···················· | ···················· | ···················· | ···················· | |

```
O 1584 hCoV-19/England/CAMC-A41018/2020|EPI_ISL_609517|20 .................... .................... ..................... .................... .................... ....................
O 1585 hCoV-19/England/CAMC-A416AD/2020|EPI_ISL_609518|20 .................... .................... ..................... .................... .................... ....................
O 1586 hCoV-19/England/CAMC-A412C7/2020|EPI_ISL_609519|20 .................... .................... ..................... .................... .................... ....................
O 1587 hCoV-19/England/CAMC-A419D1/2020|EPI_ISL_609520|20 .................... .................... ..................... .................... .................... ....................
O 1588 hCoV-19/England/CAMC-A41106/2020|EPI_ISL_609522|20 .................... .................... ..................... .................... .................... ....................
O 1589 hCoV-19/England/CAMC-A41AA1/2020|EPI_ISL_609523|20 .................... .................... ..................... .................... .................... ....................
O 1590 hCoV-19/England/CAMC-A41634/2020|EPI_ISL_609524|20 .................... .A.................. ..................... .................... .................... ..A.................
O 1591 hCoV-19/England/CAMC-A40F3A/2020|EPI_ISL_609525|20 .................... .................... ..................... .................... .................... ....................
O 1592 hCoV-19/England/CAMC-A419E0/2020|EPI_ISL_609526|20 .................... .................... ..................... .................... .................... ....................
O 1593 hCoV-19/England/CAMC-A41977/2020|EPI_ISL_609528|20 .................... .................... ..................... .................... .................... ....................
O 1594 hCoV-19/England/CAMC-A40E00/2020|EPI_ISL_609529|20 .................... .................... ..................... .................... .................... ....................
O 1595 hCoV-19/England/CAMC-A40E4C/2020|EPI_ISL_609530|20 .................... .................... ..................... .................... .................... ....................
O 1596 hCoV-19/England/CAMC-A411F7/2020|EPI_ISL_609531|20 .................... .................... ..................... .................... .................... ....................
O 1597 hCoV-19/England/CAMC-A41A29/2020|EPI_ISL_609532|20 .................... .................... ..................... .................... .................... ....................
O 1598 hCoV-19/England/CAMC-A42008/2020|EPI_ISL_609533|20 .................... .................... ..................... .................... .................... ....................
O 1599 hCoV-19/England/CAMC-A41D11/2020|EPI_ISL_609535|20 .................... .................... ..................... .................... .................... ....................
O 1600 hCoV-19/England/CAMC-A3EFA8/2020|EPI_ISL_609536|20 .................... .................... ..................... .................... .................... ....................
O 1601 hCoV-19/England/CAMC-A3EB1D/2020|EPI_ISL_609537|20 .................... .................... ..................... .................... .................... ....................
O 1602 hCoV-19/England/CAMC-A3F921/2020|EPI_ISL_609538|20 .................... .................... ..................... .................... .................... ....................
O 1603 hCoV-19/England/CAMC-A3F578/2020|EPI_ISL_609539|20 .................... .................... ..................... .................... .................... ....................
O 1604 hCoV-19/England/CAMC-A3F9F4/2020|EPI_ISL_609540|20 .................... .................... ..................... .................... .................... ....................
O 1605 hCoV-19/England/CAMC-A3EBE0/2020|EPI_ISL_609541|20 .................... .................... ..................... .................... .................... ....................
O 1606 hCoV-19/England/CAMC-A3EA89/2020|EPI_ISL_609542|20 .................... .................... ..................... .................... .................... ....................
O 1607 hCoV-19/England/CAMC-A3E62B/2020|EPI_ISL_609543|20 .................... .................... ..................... .................... .................... ....................
O 1608 hCoV-19/England/CAMC-A3F569/2020|EPI_ISL_609546|20 .................... .A.................. ..................... .................... .................... ....................
O 1609 hCoV-19/England/CAMC-A3F94F/2020|EPI_ISL_609547|20 .................... .................... ..................... .................... .................... ....................
O 1610 hCoV-19/England/CAMC-A3EEC9/2020|EPI_ISL_609548|20 .................... .................... ..................... .................... .................... ....................
O 1611 hCoV-19/England/CAMC-A3EB59/2020|EPI_ISL_609549|20 .................... .................... ..................... .................... .................... ....................
O 1612 hCoV-19/England/CAMC-A3E96E/2020|EPI_ISL_609550|20 .................... .................... ..................... .................... .................... ....................
O 1613 hCoV-19/England/CAMC-A3F88E/2020|EPI_ISL_609551|20 .................... .................... ..................... .................... .................... ....................
O 1614 hCoV-19/England/CAMC-A3F9A9/2020|EPI_ISL_609552|20 .................... .................... ..................... .................... .................... ....................
O 1615 hCoV-19/England/CAMC-A3F53C/2020|EPI_ISL_609553|20 .................... .................... ..................... .................... .................... ....................
O 1616 hCoV-19/England/CAMC-A3EA4D/2020|EPI_ISL_609554|20 .................... .................... ..................... .................... .................... ....................
O 1617 hCoV-19/England/CAMC-A3F332/2020|EPI_ISL_609555|20 .................... .................... ..................... .................... .................... ....................
O 1618 hCoV-19/England/CAMC-A3F2AE/2020|EPI_ISL_609556|20 .................... .................... ..................... .................... .................... ....................
O 1619 hCoV-19/England/CAMC-A3EDCC/2020|EPI_ISL_609557|20 .................... .................... ..................... .................... .................... ....................
O 1620 hCoV-19/England/CAMC-A3F772/2020|EPI_ISL_609558|20 .................... .................... ..................... .................... .................... ....................
O 1621 hCoV-19/England/CAMC-A3E9D7/2020|EPI_ISL_609559|20 .................... .A.................. ..................... .................... .................... ..A.................
O 1622 hCoV-19/England/CAMC-A3EE14/2020|EPI_ISL_609560|20 .................... .................... ..................... .................... .................... ....................
O 1623 hCoV-19/England/CAMC-A3EA10/2020|EPI_ISL_609561|20 .................... .................... ...........Y......... .................... .................... ....................
O 1624 hCoV-19/England/CAMC-A3E746/2020|EPI_ISL_609562|20 .................... .................... ..................... .................... .................... ....................
O 1625 hCoV-19/England/CAMC-A3E6FE/2020|EPI_ISL_609564|20 .................... .................... ..................... .................... .................... ....................
O 1626 hCoV-19/England/CAMC-A3E6D0/2020|EPI_ISL_609565|20 .................... .A.................. ..................... .................... .................... ..A.................
O 1627 hCoV-19/England/CAMC-A3EB95/2020|EPI_ISL_609566|20 .................... .A.................. ..................... .................... .................... ....................
O 1628 hCoV-19/England/CAMC-A3F244/2020|EPI_ISL_609567|20 .................... .................... ..................... .................... .................... ....................
O 1629 hCoV-19/England/CAMC-A3F87F/2020|EPI_ISL_609568|20 .................... .................... ..................... .................... .................... ....................
O 1630 hCoV-19/England/CAMC-A3F587/2020|EPI_ISL_609569|20 .................... .................... ..................... .................... .................... ....................
O 1631 hCoV-19/England/CAMC-A3F8AC/2020|EPI_ISL_609601|20 .................... .................... ..................... .................... .................... ....................
O 1632 hCoV-19/England/CAMC-A3F790/2020|EPI_ISL_609603|20 .................... .................... ..................... .................... .................... ....................
O 1633 hCoV-19/England/CAMC-A3F4D5/2020|EPI_ISL_609604|20 .................... .................... ..................... .................... .................... ....................
O 1634 hCoV-19/England/CAMC-A3F89D/2020|EPI_ISL_609605|20 .................... .................... ..................... .................... .................... ....................
O 1635 hCoV-19/England/CAMC-A3E737/2020|EPI_ISL_609606|20 .................... .A.................. ..................... .................... .................... ..A.................
O 1636 hCoV-19/England/CAMC-A3EC38/2020|EPI_ISL_609607|20 .................... .................... ..................... .................... .................... ....................
O 1637 hCoV-19/England/CAMC-A3F29F/2020|EPI_ISL_609608|20 .................... .................... ..................... .................... .................... ....................
O 1638 hCoV-19/England/CAMC-A3F754/2020|EPI_ISL_609609|20 .................... .................... ..................... .................... ......T............. ....................
O 1639 hCoV-19/England/ALDP-A85DF3/2020|EPI_ISL_610500|20 .................... .................... ..................... .................... .................... ....................
O 1640 hCoV-19/England/ALDP-A856BC/2020|EPI_ISL_610501|20 .................... .................... ..................... .................... .................... ....................
O 1641 hCoV-19/England/ALDP-A85F39/2020|EPI_ISL_610504|20 .................... .................... ..................... .................... .................... ....................
O 1642 hCoV-19/England/ALDP-A84E6A/2020|EPI_ISL_610505|20 .................... .................... ..................... .................... .................... ....................
O 1643 hCoV-19/England/ALDP-A8525E/2020|EPI_ISL_610506|20 .................... .................... ..................... .................... .................... ....................
O 1644 hCoV-19/England/ALDP-A8542B/2020|EPI_ISL_610507|20 .................... .................... ..................... .................... .................... ....................
O 1645 hCoV-19/England/ALDP-A8534C/2020|EPI_ISL_610508|20 .................... .................... ..................... .................... .................... ....................
O 1646 hCoV-19/England/ALDP-A85582/2020|EPI_ISL_610510|20 .................... .................... ..................... .................... .................... ..A.................
O 1647 hCoV-19/England/ALDP-A85F48/2020|EPI_ISL_610511|20 .................... .................... ..................... .................... .................... ....................
O 1648 hCoV-19/England/ALDP-A85CF6/2020|EPI_ISL_610513|20 .................... .................... ..................... .................... .................... ....................
O 1649 hCoV-19/England/ALDP-A84FB2/2020|EPI_ISL_610514|20 .................... .................... ..................... .................... .................... ....................
O 1650 hCoV-19/England/ALDP-A8491E/2020|EPI_ISL_610515|20 .................... .................... ..................... .................... .................... ....................
O 1651 hCoV-19/England/ALDP-A8587A/2020|EPI_ISL_610516|20 .................... .................... ..................... .................... .................... ....................
O 1652 hCoV-19/England/ALDP-A84F85/2020|EPI_ISL_610517|20 .................... .................... ..................... .................... .................... ....................
O 1653 hCoV-19/England/ALDP-A84E4C/2020|EPI_ISL_610518|20 .................... .................... ..................... .................... .................... ....................
O 1654 hCoV-19/England/ALDP-A84B72/2020|EPI_ISL_610520|20 .................... .................... ..................... .................... .................... ....................
O 1655 hCoV-19/England/ALDP-A850BE/2020|EPI_ISL_610521|20 .................... .................... ..................... .................... .................... ....................
O 1656 hCoV-19/England/ALDP-A85D3F/2020|EPI_ISL_610522|20 .................... .................... ..................... .................... .................... ....................
O 1657 hCoV-19/England/ALDP-A84C51/2020|EPI_ISL_610524|20 .................... .................... ..................... .................... .................... ....................
O 1658 hCoV-19/England/ALDP-A849D2/2020|EPI_ISL_610525|20 .................... .................... ..................... .................... .................... ....................
O 1659 hCoV-19/England/ALDP-A85B71/2020|EPI_ISL_610526|20 .................... .................... ..................... .................... .................... ....................
O 1660 hCoV-19/England/ALDP-A856AD/2020|EPI_ISL_610527|20 .................... .................... ..................... .................... .................... ....................
O 1661 hCoV-19/England/ALDP-A8527C/2020|EPI_ISL_610528|20 .................... .................... ..................... .................... .................... ....................
O 1662 hCoV-19/England/ALDP-A85221/2020|EPI_ISL_610529|20 .................... .................... ..................... .................... .................... ....................
O 1663 hCoV-19/England/ALDP-A85F75/2020|EPI_ISL_610531|20 .................... .................... ..................... .................... .................... ....................
```

| | | | | | | | |
|---|---|---|---|---|---|---|---|
| O | 1664 hCoV-19/England/ALDP-A85704/2020\|EPI_ISL_610532\|20 | ···················· | ···················· | ···················· | ···················· | ···················· | ···················· |
| O | 1665 hCoV-19/England/ALDP-A85A1A/2020\|EPI_ISL_610534\|20 | ···················· | ···················· | ···················· | ···················· | ···················· | ···················· |
| O | 1666 hCoV-19/England/ALDP-A85D11/2020\|EPI_ISL_610535\|20 | ···················· | ···················· | ···················· | ···················· | ···················· | ···················· |
| O | 1667 hCoV-19/England/ALDP-A85CC9/2020\|EPI_ISL_610536\|20 | ···················· | ···················· | ···················· | ···················· | ···················· | ···················· |
| O | 1668 hCoV-19/England/ALDP-A858F2/2020\|EPI_ISL_610538\|20 | ···················· | ···················· | ···················· | ···················· | ···················· | ···················· |
| O | 1669 hCoV-19/England/ALDP-A8528B/2020\|EPI_ISL_610539\|20 | ···················· | ···················· | ···················· | ···················· | ···················· | ···················· |
| O | 1670 hCoV-19/England/ALDP-A85519/2020\|EPI_ISL_610540\|20 | ···················· | ···················· | ···················· | ···················· | ···················· | ···················· |
| O | 1671 hCoV-19/England/ALDP-A84F1C/2020\|EPI_ISL_610541\|20 | ···················· | ···················· | ···················· | ···················· | ···················· | ···················· |
| O | 1672 hCoV-19/England/ALDP-A84D9A/2020\|EPI_ISL_610542\|20 | ···················· | ···················· | ···················· | ···················· | ···················· | ···················· |
| O | 1673 hCoV-19/England/ALDP-A854B2/2020\|EPI_ISL_610543\|20 | ···················· | ···················· | ···················· | ···················· | ···················· | ···················· |
| O | 1674 hCoV-19/England/ALDP-A851CA/2020\|EPI_ISL_610544\|20 | ···················· | ···················· | ···················· | ···················· | ···················· | ···················· |
| O | 1675 hCoV-19/England/ALDP-A85ADE/2020\|EPI_ISL_610545\|20 | ···················· | ···················· | ···················· | ···················· | ···················· | ···················· |
| O | 1676 hCoV-19/England/ALDP-A85B44/2020\|EPI_ISL_610546\|20 | ···················· | ···················· | ···················· | ···················· | ···················· | ···················· |
| O | 1677 hCoV-19/England/ALDP-A85AB0/2020\|EPI_ISL_610547\|20 | ···················· | ···················· | ···················· | ···················· | ···················· | ···················· |
| O | 1678 hCoV-19/Northern Ireland/ALDP-A8531F/2020\|EPI_ISL_ | ···················· | ···················· | ···················· | ···················· | ···················· | ···················· |
| O | 1679 hCoV-19/England/ALDP-A85CD8/2020\|EPI_ISL_610549\|20 | ···················· | ···················· | ···················· | ···················· | ···················· | ···················· |
| O | 1680 hCoV-19/England/ALDP-A85B26/2020\|EPI_ISL_610550\|20 | ···················· | ···················· | ···················· | ···················· | ···················· | ···················· |
| O | 1681 hCoV-19/England/ALDP-A84F67/2020\|EPI_ISL_610551\|20 | ···················· | ···················· | ···················· | ···················· | ···················· | ···················· |
| O | 1682 hCoV-19/England/ALDP-A84D8B/2020\|EPI_ISL_610552\|20 | ···················· | ···················· | ···················· | ···················· | ···················· | ···················· |
| O | 1683 hCoV-19/England/ALDP-A8529A/2020\|EPI_ISL_610553\|20 | ···················· | ···················· | ···················· | ···················· | ···················· | ···················· |
| O | 1684 hCoV-19/England/ALDP-A850CD/2020\|EPI_ISL_610554\|20 | ···················· | ···················· | ···················· | ···················· | ···················· | ···················· |
| O | 1685 hCoV-19/England/ALDP-A85A0B/2020\|EPI_ISL_610555\|20 | ···················· | ···················· | ···················· | ···················· | ···················· | ···················· |
| O | 1686 hCoV-19/England/ALDP-A926F8/2020\|EPI_ISL_611118\|20 | ···················· | ···················· | ···················· | ···················· | ···················· | ···················· |
| O | 1687 hCoV-19/England/EXET-138569/2020\|EPI_ISL_612400\|20 | ···················· | ···················· | ···················· | ···················· | ···················· | ···················· |
| O | 1688 hCoV-19/England/LIVE-1D29A1/2020\|EPI_ISL_612401\|20 | ···················· | ···················· | ···················· | ···················· | ···················· | ···················· |
| O | 1689 hCoV-19/England/LIVE-1D2A08/2020\|EPI_ISL_612402\|20 | ···················· | ···················· | ···················· | ···················· | ···················· | ···················· |
| O | 1690 hCoV-19/England/LIVE-1D2A71/2020\|EPI_ISL_612403\|20 | ···················· | ···················· | ···················· | ···················· | ···················· | ···················· |
| O | 1692 hCoV-19/England/LIVE-1D2AAE/2020\|EPI_ISL_612405\|20 | ···················· | ···················· | ···················· | ···················· | ···················· | ···················· |
| O | 1694 hCoV-19/England/LIVE-1D2AF9/2020\|EPI_ISL_612407\|20 | ···················· | ···················· | ···················· | ···················· | ···················· | ···················· |
| O | 1695 hCoV-19/England/LIVE-1D2B23/2020\|EPI_ISL_612408\|20 | ···················· | ···················· | ···················· | ···················· | ···················· | ···················· |
| O | 1696 hCoV-19/England/LIVE-1D2B41/2020\|EPI_ISL_612409\|20 | ···················· | ···················· | ···················· | ···················· | ···················· | ···················· |
| O | 1697 hCoV-19/England/LIVE-1D2B7E/2020\|EPI_ISL_612410\|20 | ···················· | ···················· | ···················· | ···················· | ···················· | ···················· |
| O | 1698 hCoV-19/England/LIVE-1D2B8D/2020\|EPI_ISL_612411\|20 | ···················· | ···················· | ···················· | ···················· | ···················· | ···················· |
| O | 1699 hCoV-19/England/LIVE-1D2B9C/2020\|EPI_ISL_612412\|20 | ···················· | ···················· | ···················· | ···················· | ···················· | ···················· |
| O | 1700 hCoV-19/England/LIVE-1D2BAB/2020\|EPI_ISL_612413\|20 | ···················· | ···················· | ···················· | ···················· | ···················· | ···················· |
| O | 1702 hCoV-19/England/LIVE-1D2C02/2020\|EPI_ISL_612415\|20 | ···················· | ···················· | ···················· | ···················· | ···················· | ···················· |
| O | 1703 hCoV-19/England/LIVE-ABA72/2020\|EPI_ISL_612449\|202 | ···················· | ···················· | ···················· | ···················· | ···················· | ···················· |
| O | 1704 hCoV-19/Gibraltar/204421244/2020\|EPI_ISL_613703\|20 | ···················· | ···················· | ···················· | ···················· | ···················· | ···················· |
| O | 1705 hCoV-19/Gibraltar/204421245/2020\|EPI_ISL_613704\|20 | ···················· | ···················· | ···················· | ···················· | ···················· | ···················· |
| O | 1706 hCoV-19/Gibraltar/204421248/2020\|EPI_ISL_613705\|20 | ···················· | ···················· | ···················· | ···················· | ···················· | ···················· |
| O | 1707 hCoV-19/Italy/SAR-ASSL-02/2020\|EPI_ISL_613706\|2020 | ···················· | ···················· | ···················· | ···················· | ···················· | ···················· |
| O | 1708 hCoV-19/Brazil/SP-RP4/2020\|EPI_ISL_613707\|2020-03- | ···················· | ···················· | ···················· | ···················· | ···················· | ···················· |
| O | 1709 hCoV-19/Brazil/SP-RP21/2020\|EPI_ISL_613708\|2020-04 | ···················· | ···················· | ···················· | ···················· | ···················· | ···················· |
| O | 1710 hCoV-19/Brazil/SP-RP2/2020\|EPI_ISL_613709\|2020-03- | ···················· | ···················· | ···················· | ···················· | ···················· | ···················· |
| O | 1711 hCoV-19/USA/VA-DCLS-1856/2020\|EPI_ISL_614080\|2020- | ···················· | ···A················ | ···················· | ···················· | ···················· | ···················· |
| O | 1712 hCoV-19/USA/VA-DCLS-1857/2020\|EPI_ISL_614081\|2020- | ···················· | ···················· | ···················· | ···················· | ···················· | ···················· |
| O | 1713 hCoV-19/USA/VA-DCLS-1860/2020\|EPI_ISL_614082\|2020- | ···················· | ···················· | ···················· | ···················· | ···················· | ···················· |
| O | 1714 hCoV-19/USA/VA-DCLS-1862/2020\|EPI_ISL_614083\|2020- | ···················· | ···················· | ···················· | ···················· | ···················· | ···················· |
| O | 1715 hCoV-19/USA/VA-DCLS-1863/2020\|EPI_ISL_614084\|2020- | ···················· | ···················· | ···················· | ···················· | ···················· | ···················· |
| O | 1716 hCoV-19/USA/VA-DCLS-1865/2020\|EPI_ISL_614085\|2020- | ···················· | ·A·················· | ···················· | ···················· | ···················· | ···················· |
| O | 1717 hCoV-19/USA/VA-DCLS-1867/2020\|EPI_ISL_614086\|2020- | ···················· | ···················· | ···················· | ···················· | ···················· | ···················· |
| O | 1718 hCoV-19/USA/VA-DCLS-1869/2020\|EPI_ISL_614087\|2020- | ···················· | ···················· | ···················· | ···················· | ···················· | ···················· |
| O | 1719 hCoV-19/USA/VA-DCLS-1870/2020\|EPI_ISL_614088\|2020- | ···················· | ·A·················· | ···················· | ···················· | ···················· | ···················· |
| O | 1720 hCoV-19/USA/VA-DCLS-1900/2020\|EPI_ISL_614089\|2020- | ···················· | ···················· | ···················· | ···················· | ···················· | ···················· |
| O | 1721 hCoV-19/USA/VA-DCLS-1902/2020\|EPI_ISL_614090\|2020- | ···················· | ···················· | ···················· | ···················· | ···················· | ···················· |
| O | 1722 hCoV-19/USA/VA-DCLS-1903/2020\|EPI_ISL_614091\|2020- | ···················· | ···················· | ···················· | ···················· | ···················· | ···················· |
| O | 1723 hCoV-19/USA/VA-DCLS-1905/2020\|EPI_ISL_614092\|2020- | ···················· | ···················· | ···················· | ···················· | ···················· | ···················· |
| O | 1724 hCoV-19/USA/VA-DCLS-1924/2020\|EPI_ISL_614093\|2020- | ···················· | ···················· | ···················· | ···················· | ···················· | ···················· |
| O | 1725 hCoV-19/USA/VA-DCLS-1985/2020\|EPI_ISL_614094\|2020- | ···················· | ·A·················· | ···················· | ···················· | ···················· | ···················· |
| O | 1726 hCoV-19/USA/VA-DCLS-1990/2020\|EPI_ISL_614096\|2020- | ···················· | ·A·················· | ···················· | ···················· | ···················· | ···················· |
| O | 1727 hCoV-19/USA/VA-DCLS-1991/2020\|EPI_ISL_614097\|2020- | ···················· | ·A·················· | ···················· | ···················· | ···················· | ···················· |
| O | 1728 hCoV-19/USA/VA-DCLS-1992/2020\|EPI_ISL_614098\|2020- | ···················· | ·A·················· | ···················· | ···················· | ···················· | ···················· |
| O | 1729 hCoV-19/USA/VA-DCLS-1993/2020\|EPI_ISL_614099\|2020- | ···················· | ·A·················· | ···················· | ···················· | ···················· | ···················· |
| O | 1730 hCoV-19/USA/VA-DCLS-0311/2020\|EPI_ISL_614100\|2020- | ···················· | ···················· | ···················· | ···················· | ···················· | ···················· |
| O | 1731 hCoV-19/USA/VA-DCLS-0312/2020\|EPI_ISL_614101\|2020- | ···················· | ···················· | ···················· | ···················· | ···················· | ···················· |
| O | 1732 hCoV-19/USA/VA-DCLS-0313/2020\|EPI_ISL_614102\|2020- | ···················· | ···················· | ···················· | ···················· | ···················· | ···················· |
| O | 1733 hCoV-19/USA/VA-DCLS-0314/2020\|EPI_ISL_614103\|2020- | ···················· | ···················· | ···················· | ···················· | ···················· | ···················· |
| O | 1734 hCoV-19/USA/VA-DCLS-0323/2020\|EPI_ISL_614104\|2020- | ···················· | ···················· | ···················· | ···················· | ···················· | ···················· |
| O | 1735 hCoV-19/USA/VA-DCLS-0324/2020\|EPI_ISL_614105\|2020- | ···················· | ···················· | ···················· | ···················· | ···················· | ···················· |
| O | 1736 hCoV-19/USA/VA-DCLS-0326/2020\|EPI_ISL_614107\|2020- | ···················· | ···················· | ···················· | ···················· | ···················· | ···················· |
| O | 1737 hCoV-19/USA/VA-DCLS-0338/2020\|EPI_ISL_614108\|2020- | ···················· | ···················· | ···················· | ···················· | ···················· | ···················· |
| O | 1738 hCoV-19/USA/VA-DCLS-0490/2020\|EPI_ISL_614109\|2020- | ···················· | ···················· | ···················· | ···················· | ···················· | ···················· |
| O | 1739 hCoV-19/USA/VA-DCLS-0491/2020\|EPI_ISL_614110\|2020- | ···················· | ···················· | ···················· | ···················· | ···················· | ···················· |
| O | 1740 hCoV-19/USA/VA-DCLS-0501/2020\|EPI_ISL_614111\|2020- | ···················· | ···················· | ···················· | ···················· | ···················· | ···················· |
| O | 1741 hCoV-19/USA/VA-DCLS-0502/2020\|EPI_ISL_614112\|2020- | ···················· | ···················· | ···················· | ···················· | ···················· | ···················· |
| O | 1742 hCoV-19/USA/VA-DCLS-0504/2020\|EPI_ISL_614113\|2020- | ···················· | ···················· | ···················· | ···················· | ···················· | ···················· |
| O | 1743 hCoV-19/USA/VA-DCLS-0505/2020\|EPI_ISL_614114\|2020- | ···················· | ···················· | ···················· | ···················· | ···················· | ···················· |
| O | 1744 hCoV-19/USA/VA-DCLS-0514/2020\|EPI_ISL_614115\|2020- | ···················· | ···················· | ···················· | ···················· | ···················· | ···················· |
| O | 1745 hCoV-19/USA/VA-DCLS-0515/2020\|EPI_ISL_614116\|2020- | ···················· | ···················· | ···················· | ···················· | ···················· | ···················· |
| O | 1746 hCoV-19/USA/VA-DCLS-0516/2020\|EPI_ISL_614117\|2020- | ···················· | ···················· | ···················· | ···················· | ···················· | ···················· |

O 1747 hCoV-19/USA/VA-DCLS-0517/2020|EPI_ISL_614118|2020- ···················· ···················· ···················· ···················· ···················· ····················
O 1748 hCoV-19/USA/VA-DCLS-0518/2020|EPI_ISL_614119|2020- ···················· ···················· ···················· ···················· ···················· ····················
O 1749 hCoV-19/USA/VA-DCLS-0519/2020|EPI_ISL_614120|2020- ···················· ···················· ···················· ···················· ···················· ····················
O 1750 hCoV-19/USA/VA-DCLS-0520/2020|EPI_ISL_614121|2020- ···················· ···················· ···················· ···················· ···················· ····················
O 1751 hCoV-19/USA/VA-DCLS-0522/2020|EPI_ISL_614122|2020- ···················· ···················· ···················· ···················· ···················· ····················
O 1752 hCoV-19/USA/VA-DCLS-1713/2020|EPI_ISL_614123|2020- ···················· ·A·················· ···················· ···················· ···················· ····················
O 1753 hCoV-19/USA/VA-DCLS-1714/2020|EPI_ISL_614124|2020- ···················· ·A·················· ···················· ···················· ···················· ····················
O 1754 hCoV-19/USA/VA-DCLS-1715/2020|EPI_ISL_614125|2020- ···················· ···················· ···················· ···················· ···················· ····················
O 1755 hCoV-19/USA/VA-DCLS-1716/2020|EPI_ISL_614126|2020- ···················· ·A·················· ···················· ···················· ···················· ····················
O 1756 hCoV-19/USA/VA-DCLS-1717/2020|EPI_ISL_614127|2020- ···················· ···················· ···················· ···················· ···················· ····················
O 1757 hCoV-19/USA/VA-DCLS-1836/2020|EPI_ISL_614128|2020- ···················· ···················· ···················· ···················· ···················· ····················
O 1758 hCoV-19/USA/VA-DCLS-1837/2020|EPI_ISL_614129|2020- ···················· ···················· ···················· ···················· ···················· ····················
O 1759 hCoV-19/USA/VA-DCLS-1838/2020|EPI_ISL_614130|2020- ···········T········ ···················· ···················· ···················· ···················· ····················
O 1760 hCoV-19/USA/VA-DCLS-1839/2020|EPI_ISL_614131|2020- ···········T········ ···················· ···················· ···················· ···················· ····················
O 1761 hCoV-19/USA/VA-DCLS-1840/2020|EPI_ISL_614132|2020- ···················· ···················· ···················· ···················· ···················· ····················
O 1762 hCoV-19/USA/VA-DCLS-1841/2020|EPI_ISL_614133|2020- ···················· ···················· ···················· ···················· ···················· ····················
O 1763 hCoV-19/USA/VA-DCLS-1842/2020|EPI_ISL_614134|2020- ···················· ···················· ···················· ···················· ···················· ····················
O 1764 hCoV-19/USA/VA-DCLS-1844/2020|EPI_ISL_614135|2020- ···················· ···················· ···················· ···················· ···················· ····················
O 1765 hCoV-19/USA/VA-DCLS-1816/2020|EPI_ISL_614136|2020- ···················· ·A·················· ···················· ···················· ···················· ····················
O 1766 hCoV-19/USA/VA-DCLS-1817/2020|EPI_ISL_614137|2020- ···················· ·A·················· ···················· ···················· ···················· ····················
O 1767 hCoV-19/USA/VA-DCLS-1818/2020|EPI_ISL_614138|2020- ···················· ·A·················· ···················· ···················· ···················· ····················
O 1768 hCoV-19/USA/VA-DCLS-1819/2020|EPI_ISL_614139|2020- ···················· ·A·················· ···················· ···················· ···················· ····················
O 1769 hCoV-19/USA/VA-DCLS-1821/2020|EPI_ISL_614140|2020- ···················· ·A·················· ···················· ···················· ···················· ····················
O 1770 hCoV-19/USA/VA-DCLS-1823/2020|EPI_ISL_614141|2020- ···················· ···················· ···················· ···················· ···················· ····················
O 1771 hCoV-19/USA/VA-DCLS-1824/2020|EPI_ISL_614142|2020- ···················· ···················· ···················· ···················· ···················· ····················
O 1772 hCoV-19/USA/VA-DCLS-1827/2020|EPI_ISL_614143|2020- ···················· ···················· ···················· ···················· ···················· ····················
O 1773 hCoV-19/USA/VA-DCLS-1829/2020|EPI_ISL_614144|2020- ···················· ···················· ···················· ···················· ···················· ····················
O 1774 hCoV-19/USA/VA-DCLS-1889/2020|EPI_ISL_614145|2020- ···················· ···················· ···················· ···················· ···················· ····················
O 1775 hCoV-19/USA/VA-DCLS-1890/2020|EPI_ISL_614146|2020- ···················· ···················· ···················· ···················· ···················· ····················
O 1776 hCoV-19/USA/VA-DCLS-1892/2020|EPI_ISL_614147|2020- ···················· ···················· ···················· ···················· ···················· ····················
O 1777 hCoV-19/USA/VA-DCLS-1893/2020|EPI_ISL_614148|2020- ···················· ···················· ···················· ···················· ···················· ····················
O 1779 hCoV-19/USA/MI-MDHHS-SC22117/2020|EPI_ISL_614168|2 ···················· ···················· ···················· ···················· ···················· ····················
O 1783 hCoV-19/USA/MI-MDHHS-SC22121/2020|EPI_ISL_614172|2 ···················· ···················· ···················· ···················· ···················· ····················
O 1784 hCoV-19/USA/MI-MDHHS-SC22122/2020|EPI_ISL_614173|2 ···················· ·A·················· ···················· ···················· ···················· ····················
O 1785 hCoV-19/USA/MI-MDHHS-SC22123/2020|EPI_ISL_614174|2 ···················· ·············A······ ···················· ···················· ···················· ····················
O 1786 hCoV-19/USA/MI-MDHHS-SC22124/2020|EPI_ISL_614175|2 ···················· ·A·················· ···················· ···················· ···················· ····················
O 1787 hCoV-19/USA/MI-MDHHS-SC22125/2020|EPI_ISL_614176|2 ···················· ··········A········· ···················· ···················· ···················· ····················
O 1788 hCoV-19/USA/MI-MDHHS-SC22126/2020|EPI_ISL_614177|2 ···················· ·A·················· ···················· ···················· ···················· ····················
O 1789 hCoV-19/USA/MI-MDHHS-SC22127/2020|EPI_ISL_614178|2 ···················· ···················· ···················· ···················· ··N················· ····················
O 1790 hCoV-19/USA/MI-MDHHS-SC22128/2020|EPI_ISL_614179|2 ···················· ·A·················· ···················· ···················· ···················· ····················
O 1791 hCoV-19/USA/MI-MDHHS-SC22129/2020|EPI_ISL_614180|2 ···················· ···················· ···················· ···················· ···················· ····················
O 1792 hCoV-19/USA/MI-MDHHS-SC22148/2020|EPI_ISL_614199|2 ···················· ···················· ···················· ···················· ···················· ····················
O 1793 hCoV-19/England/ALDP-A8730E/2020|EPI_ISL_605932|20 ···················· ···················· ···················· ···················· ···················· ····················
O 1794 hCoV-19/England/ALDP-A86DB6/2020|EPI_ISL_605933|20 ···················· ···················· ···················· ···················· ···················· ····················
O 1795 hCoV-19/England/ALDP-A87B9D/2020|EPI_ISL_605934|20 ···················· ···················· ···················· ···················· ···················· ····················
O 1796 hCoV-19/England/ALDP-A87580/2020|EPI_ISL_605936|20 ···················· ···················· ···················· ···················· ···················· ····················
O 1797 hCoV-19/England/ALDP-A87939/2020|EPI_ISL_605937|20 ···················· ···················· ···················· ···················· ···················· ····················
O 1798 hCoV-19/England/ALDP-A86ED1/2020|EPI_ISL_605939|20 ···················· ···················· ···················· ···················· ···················· ····················
O 1799 hCoV-19/England/ALDP-A87562/2020|EPI_ISL_605940|20 ···················· ···················· ···················· ···················· ···················· ····················
O 1800 hCoV-19/England/ALDP-A87D97/2020|EPI_ISL_605941|20 ···················· ···················· ···················· ···················· ···················· ····················
O 1801 hCoV-19/England/ALDP-A87C4F/2020|EPI_ISL_605942|20 ···················· ···················· ···················· ···················· ······T············· ····················
O 1802 hCoV-19/England/ALDP-A87984/2020|EPI_ISL_605943|20 ···················· ···················· ···················· ···················· ···················· ····················
O 1803 hCoV-19/England/ALDP-A8641B/2020|EPI_ISL_605945|20 ···················· ···················· ···················· ···················· ···················· ····················
O 1804 hCoV-19/England/ALDP-A86F1A/2020|EPI_ISL_605947|20 ···················· ···················· ···················· ···················· ····T··············· ····················
O 1805 hCoV-19/England/ALDP-A861E7/2020|EPI_ISL_605948|20 ···················· ···················· ···················· ···················· ···················· ····················
O 1806 hCoV-19/England/ALDP-A8634B/2020|EPI_ISL_605949|20 ···················· ···················· ···················· ···················· ···················· ····················
O 1807 hCoV-19/England/ALDP-A8668E/2020|EPI_ISL_605950|20 ···················· ···················· ···················· ···················· ···················· ····················
O 1808 hCoV-19/England/ALDP-A86FA1/2020|EPI_ISL_605951|20 ···················· ···················· ···················· ···················· ···················· ····················
O 1809 hCoV-19/England/ALDP-A87016/2020|EPI_ISL_605952|20 ···················· ···················· ···················· ···················· ···················· ····················
O 1810 hCoV-19/England/ALDP-A87025/2020|EPI_ISL_605953|20 ···················· ···················· ···················· ···················· ···················· ····················
O 1811 hCoV-19/England/ALDP-A86518/2020|EPI_ISL_605954|20 ···················· ·A·················· ···················· ···················· ···················· ····················
O 1812 hCoV-19/England/ALDP-A86C9B/2020|EPI_ISL_605955|20 ···················· ···················· ···················· ···················· ···················· ····················
O 1813 hCoV-19/England/ALDP-A864DF/2020|EPI_ISL_605956|20 ···················· ···················· ···················· ···················· ···················· ····················
O 1814 hCoV-19/England/ALDP-A870DA/2020|EPI_ISL_605959|20 ···················· ···················· ···················· ···················· ···················· ····················
O 1815 hCoV-19/England/ALDP-A86660/2020|EPI_ISL_605960|20 ···················· ···················· ···················· ···················· ···················· ····················
O 1816 hCoV-19/England/ALDP-A87799/2020|EPI_ISL_605961|20 ···················· ···················· ···················· ···················· ···················· ····················
O 1817 hCoV-19/England/ALDP-A864B1/2020|EPI_ISL_605962|20 ···················· ···················· ···················· ···················· ···················· ····················
O 1818 hCoV-19/England/ALDP-A864A2/2020|EPI_ISL_605965|20 ···················· ···················· ···················· ···················· ···················· ····················
O 1819 hCoV-19/England/ALDP-A87465/2020|EPI_ISL_605966|20 ···················· ···················· ···················· ···················· ···················· ····················
O 1820 hCoV-19/England/ALDP-A86DE3/2020|EPI_ISL_605967|20 ···················· ···················· ···················· ···················· ···················· ····················
O 1821 hCoV-19/England/ALDP-A871C8/2020|EPI_ISL_605968|20 ···················· ···················· ···················· ···················· ···················· ····················
O 1822 hCoV-19/England/ALDP-A87ABE/2020|EPI_ISL_605969|20 ···················· ···················· ···················· ···················· ···················· ····················
O 1823 hCoV-19/England/ALDP-A87650/2020|EPI_ISL_605970|20 ···················· ···················· ···················· ···················· ···················· ····················
O 1824 hCoV-19/England/ALDP-A8726B/2020|EPI_ISL_605971|20 ···················· ···················· ···················· ···················· ···················· ····················
O 1825 hCoV-19/England/ALDP-A877A8/2020|EPI_ISL_605972|20 ···················· ···················· ···················· ···················· ···················· ····················
O 1826 hCoV-19/England/ALDP-A87CF4/2020|EPI_ISL_605973|20 ···················· ···················· ···················· ···················· ···················· ····················
O 1827 hCoV-19/England/ALDP-A87571/2020|EPI_ISL_605975|20 ···················· ···················· ···················· ···················· ···················· ····················
O 1828 hCoV-19/England/ALDP-A872E3/2020|EPI_ISL_605976|20 ···················· ···················· ···················· ···················· ···················· ····················
O 1829 hCoV-19/England/ALDP-A8628A/2020|EPI_ISL_605977|20 ···················· ···················· ···················· ···················· ···················· ····················
O 1830 hCoV-19/England/ALDP-A86C40/2020|EPI_ISL_605978|20 ···················· ···················· ···················· ···················· ···················· ····················

O 1831 hCoV-19/England/ALDP-A86E59/2020|EPI_ISL_605980|20 ···················· ···················· ···················· ···················· ···················· ····················
O 1832 hCoV-19/England/ALDP-A87377/2020|EPI_ISL_605981|20 ···················· ···················· ···················· ···················· ···················· ····················
O 1833 hCoV-19/England/ALDP-A877F3/2020|EPI_ISL_605982|20 ···················· ···················· ···················· ···················· ···················· ····················
O 1835 hCoV-19/England/ALDP-A86466/2020|EPI_ISL_605984|20 ···················· ···················· ···················· ···················· ···················· ····················
O 1836 hCoV-19/England/ALDP-A86536/2020|EPI_ISL_605985|20 ···················· ···················· ···················· ···················· ···················· ····················
O 1837 hCoV-19/England/ALDP-A86299/2020|EPI_ISL_605986|20 ···················· ···················· ···················· ···················· ···················· ····················
O 1838 hCoV-19/England/ALDP-A87492/2020|EPI_ISL_605987|20 ···················· ···················· ···················· ···················· ···················· ····················
O 1839 hCoV-19/England/ALDP-A86114/2020|EPI_ISL_605989|20 ···················· ···················· ···················· ···················· ···················· ····················
O 1840 hCoV-19/England/ALDP-A870BC/2020|EPI_ISL_605991|20 ···················· ···················· ···················· ···················· ···················· ···A················
O 1841 hCoV-19/England/ALDP-A877D5/2020|EPI_ISL_605992|20 ···················· ·A·················· ···················· ···················· ···················· ····················
O 1842 hCoV-19/England/ALDP-A87DA6/2020|EPI_ISL_605993|20 ···················· ···················· ···················· ···················· ···················· ····················
O 1843 hCoV-19/England/ALDP-A8717D/2020|EPI_ISL_605994|20 ···················· ···················· ···················· ···················· ···················· ····················
O 1844 hCoV-19/England/ALDP-A87043/2020|EPI_ISL_605995|20 ···················· ···················· ···················· ···················· ···················· ····················
O 1845 hCoV-19/England/ALDP-A6CD6B/2020|EPI_ISL_608240|20 ···················· ···················· ···················· ···················· ···················· ····················
O 1846 hCoV-19/England/ALDP-A6D526/2020|EPI_ISL_608241|20 ···················· ···················· ···················· ···················· ···················· ····················
O 1847 hCoV-19/England/ALDP-A6CB52/2020|EPI_ISL_608243|20 ···················· ···················· ···················· ···················· ···················· ····················
O 1848 hCoV-19/England/ALDP-A6CB9E/2020|EPI_ISL_608244|20 ···················· ···················· ···················· ···················· ···················· ····················
O 1849 hCoV-19/England/ALDP-A6CBBC/2020|EPI_ISL_608245|20 ···················· ···················· ···················· ···················· ···················· ····················
O 1850 hCoV-19/England/ALDP-A6FB04/2020|EPI_ISL_608246|20 ···················· ···················· ···················· ···················· ···················· ····················
O 1851 hCoV-19/England/ALDP-A6D1D7/2020|EPI_ISL_608247|20 ···················· ···················· ···················· ···················· ···················· ····················
O 1852 hCoV-19/England/ALDP-A707C0/2020|EPI_ISL_608248|20 ···················· ···················· ···················· ···················· ···················· ····················
O 1853 hCoV-19/England/ALDP-A6D456/2020|EPI_ISL_608249|20 ···················· ···················· ···················· ···················· ···················· ····················
O 1854 hCoV-19/England/ALDP-A70818/2020|EPI_ISL_608250|20 ···················· ···················· ···················· ···················· ···················· ····················
O 1855 hCoV-19/England/ALDP-A6D1E6/2020|EPI_ISL_608253|20 ···················· ···················· ···················· ···················· ···················· ····················
O 1856 hCoV-19/England/ALDP-A6D30E/2020|EPI_ISL_608254|20 ···················· ···················· ···················· ···················· ···················· ····················
O 1857 hCoV-19/England/ALDP-A6FF17/2020|EPI_ISL_608255|20 ···················· ·A·················· ···················· ···················· ···················· ···A················
O 1858 hCoV-19/England/ALDP-A6D8E1/2020|EPI_ISL_608256|20 ···················· ···················· ···················· ···················· ···················· ····················
O 1859 hCoV-19/England/ALDP-A70784/2020|EPI_ISL_608257|20 ···················· ···················· ···················· ···················· ···················· ····················
O 1860 hCoV-19/England/ALDP-A6CC9B/2020|EPI_ISL_608259|20 ···················· ···················· ···················· ···················· ···················· ····················
O 1861 hCoV-19/England/ALDP-A6CDF2/2020|EPI_ISL_608260|20 ···················· ···················· ···················· ···················· ···················· ····················
O 1862 hCoV-19/England/ALDP-A6C68E/2020|EPI_ISL_608261|20 ···················· ···················· ···················· ···················· ···················· ····················
O 1863 hCoV-19/England/ALDP-A70827/2020|EPI_ISL_608262|20 ···················· ···················· ···················· ···················· ···················· ····················
O 1864 hCoV-19/England/ALDP-A6FE38/2020|EPI_ISL_608263|20 ···················· ·A·················· ···················· ···················· ···················· ····················
O 1865 hCoV-19/England/ALDP-A6C879/2020|EPI_ISL_608264|20 ···················· ···················· ···················· ···················· ···················· ····················
O 1866 hCoV-19/England/ALDP-A70951/2020|EPI_ISL_608265|20 ···················· ···················· ···················· ···················· ···················· ····················
O 1867 hCoV-19/England/ALDP-A6C4C0/2020|EPI_ISL_608266|20 ···················· ·A·················· ···················· ···················· ···················· ···A················
O 1868 hCoV-19/England/ALDP-A6D78A/2020|EPI_ISL_608267|20 ···················· ···················· ···················· ···················· ···················· ····················
O 1869 hCoV-19/England/ALDP-A6D73F/2020|EPI_ISL_608268|20 ···················· ···················· ···················· ···················· ···················· ····················
O 1870 hCoV-19/England/ALDP-A6C800/2020|EPI_ISL_608269|20 ···················· ···················· ···················· ···················· ···················· ····················
O 1871 hCoV-19/England/ALDP-A6CFA1/2020|EPI_ISL_608270|20 ···················· ···················· ···················· ···················· ···················· ····················
O 1872 hCoV-19/England/ALDP-A6C6AC/2020|EPI_ISL_608271|20 ···················· ···················· ···················· ···················· ···················· ····················
O 1873 hCoV-19/England/ALDP-A6CEB3/2020|EPI_ISL_608272|20 ···················· ···················· ···················· ···················· ···················· ····················
O 1874 hCoV-19/England/ALDP-A6C42A/2020|EPI_ISL_608273|20 ···················· ···················· ···················· ···················· ···················· ····················
O 1875 hCoV-19/England/ALDP-A6C4B1/2020|EPI_ISL_608274|20 ···················· ···················· ···················· ···················· ···················· ····················
O 1876 hCoV-19/England/ALDP-A6C563/2020|EPI_ISL_608275|20 ···················· ···················· ···················· ···················· ···················· ····················
O 1877 hCoV-19/England/ALDP-A6D2A7/2020|EPI_ISL_608276|20 ···················· ···················· ···················· ···················· ···················· ····················
O 1878 hCoV-19/England/ALDP-A6D377/2020|EPI_ISL_608277|20 ···················· ···················· ···················· ···················· ···················· ····················
O 1879 hCoV-19/England/ALDP-A7012C/2020|EPI_ISL_608278|20 ···················· ···················· ···················· ···················· ···················· ····················
O 1880 hCoV-19/England/ALDP-A70247/2020|EPI_ISL_608279|20 ···················· ···················· ···················· ···················· ···················· ····················
O 1881 hCoV-19/England/ALDP-A6FFBD/2020|EPI_ISL_608280|20 ···················· ···················· ···················· ···················· ···················· ····················
O 1882 hCoV-19/England/ALDP-A7004D/2020|EPI_ISL_608281|20 ···················· ···················· ···················· ···················· ···················· ····················
O 1883 hCoV-19/England/ALDP-A6C651/2020|EPI_ISL_608282|20 ···················· ···················· ···················· ···················· ···················· ····················
O 1884 hCoV-19/England/ALDP-A6D395/2020|EPI_ISL_608283|20 ···················· ···················· ···················· ···················· ···················· ····················
O 1885 hCoV-19/England/ALDP-A6CD01/2020|EPI_ISL_608286|20 ···················· ···················· ···················· ···················· ···················· ····················
O 1886 hCoV-19/England/ALDP-A70177/2020|EPI_ISL_608287|20 ···················· ···················· ···················· ···················· ···················· ····················
O 1887 hCoV-19/England/ALDP-A6D2C5/2020|EPI_ISL_608288|20 ···················· ···················· ···················· ···················· ···················· ····················
O 1888 hCoV-19/England/ALDP-A6D3A4/2020|EPI_ISL_608289|20 ···················· ···················· ···················· ···················· ···················· ····················
O 1889 hCoV-19/England/ALDP-A6C7F4/2020|EPI_ISL_608291|20 ···················· ···················· ···················· ···················· ···················· ····················
O 1890 hCoV-19/England/ALDP-A6D438/2020|EPI_ISL_608292|20 ···················· ···················· ···················· ···················· ···················· ····················
O 1891 hCoV-19/England/ALDP-A6FFAE/2020|EPI_ISL_608293|20 ···················· ···················· ···················· ···················· ···················· ····················
O 1892 hCoV-19/England/ALDP-A6D0BC/2020|EPI_ISL_608296|20 ···················· ···················· ···················· ···················· ···················· ····················
O 1893 hCoV-19/England/ALDP-A6D0F8/2020|EPI_ISL_608298|20 ···················· ···················· ···················· ···················· ···················· ····················
O 1894 hCoV-19/England/ALDP-A6C606/2020|EPI_ISL_608299|20 ···················· ···················· ···················· ···················· ···················· ····················
O 1895 hCoV-19/England/CAMC-A41C05/2020|EPI_ISL_609410|20 ···················· ···················· ···················· ···················· ···················· ····················
O 1896 hCoV-19/England/CAMC-A41036/2020|EPI_ISL_609411|20 ···················· ···················· ···················· ···················· ···················· ····················
O 1897 hCoV-19/England/CAMC-A4194A/2020|EPI_ISL_609412|20 ···················· ···················· ···················· ···················· ···················· ····················
O 1898 hCoV-19/England/CAMC-A41CF6/2020|EPI_ISL_609413|20 ···················· ···················· ···················· ···················· ···················· ····················
O 1899 hCoV-19/England/CAMC-A41A74/2020|EPI_ISL_609414|20 ···················· ···················· ···················· ···················· ···················· ····················
O 1900 hCoV-19/England/CAMC-A42080/2020|EPI_ISL_609415|20 ···················· ···················· ···················· ···················· ···················· ····················
O 1901 hCoV-19/England/CAMC-A40F67/2020|EPI_ISL_609416|20 ···················· ···················· ···················· ···················· ···················· ····················
O 1902 hCoV-19/England/CAMC-A41E87/2020|EPI_ISL_609417|20 ···················· ···················· ···················· ···················· ···················· ····················
O 1903 hCoV-19/England/CAMC-A40EF1/2020|EPI_ISL_609418|20 ···················· ···················· ···················· ···················· ···················· ····················
O 1904 hCoV-19/England/CAMC-A41203/2020|EPI_ISL_609423|20 ···················· ···················· ···················· ···················· ···················· ····················
O 1905 hCoV-19/England/CAMC-A4141C/2020|EPI_ISL_609424|20 ···················· ···················· ···················· ···················· ···················· ····················
O 1906 hCoV-19/England/CAMC-A40F0D/2020|EPI_ISL_609426|20 ···················· ···················· ···················· ···················· ···················· ····················
O 1907 hCoV-19/England/CAMC-A41A38/2020|EPI_ISL_609427|20 ···················· ···················· ···················· ···················· ······T············· ····················
O 1908 hCoV-19/England/CAMC-A41054/2020|EPI_ISL_609428|20 ···················· ···················· ···················· ···················· ···················· ····················
O 1909 hCoV-19/England/CAMC-A413D3/2020|EPI_ISL_609429|20 ···················· ···················· ···················· ···················· ···················· ····················
O 1910 hCoV-19/England/CAMC-A3F5B4/2020|EPI_ISL_609570|20 ···················· ···················· ···················· ···················· ···················· ····················
O 1911 hCoV-19/England/CAMC-A3F9D6/2020|EPI_ISL_609571|20 ···················· ···················· ···················· ···················· ···················· ····················

| | | | | | | | | |
|---|---|---|---|---|---|---|---|---|
| O | 1912 | hCoV-19/England/CAMC-A3E3E8/2020\|EPI_ISL_609572\|20 | ···················· | ···················· | ····················· | ···················· | ···················· | ···················· |
| O | 1913 | hCoV-19/England/CAMC-A3ECA1/2020\|EPI_ISL_609573\|20 | ···················· | ·A·················· | ····················· | ···················· | ···················· | ···A················ |
| O | 1914 | hCoV-19/England/CAMC-A3E8BC/2020\|EPI_ISL_609574\|20 | ···················· | ···················· | ····················· | ···················· | ···················· | ···················· |
| O | 1915 | hCoV-19/England/CAMC-A3FA6A/2020\|EPI_ISL_609575\|20 | ···················· | ···················· | ····················· | ···················· | ···················· | ···················· |
| O | 1916 | hCoV-19/England/CAMC-A3F52D/2020\|EPI_ISL_609576\|20 | ···················· | ···················· | ····················· | ···················· | ···················· | ···················· |
| O | 1917 | hCoV-19/England/CAMC-A3F7CD/2020\|EPI_ISL_609577\|20 | ···················· | ···················· | ····················· | ···················· | ···················· | ···················· |
| O | 1918 | hCoV-19/England/CAMC-A3EF99/2020\|EPI_ISL_609578\|20 | ···················· | ···················· | ····················· | ···················· | ···················· | ···················· |
| O | 1919 | hCoV-19/England/CAMC-A3F4A8/2020\|EPI_ISL_609579\|20 | ···················· | ···················· | ····················· | ···················· | ···················· | ···················· |
| O | 1920 | hCoV-19/England/CAMC-A3F0C2/2020\|EPI_ISL_609580\|20 | ···················· | ···················· | ····················· | ···················· | ···················· | ···················· |
| O | 1921 | hCoV-19/England/CAMC-A3F684/2020\|EPI_ISL_609582\|20 | ···················· | ···················· | ····················· | ···················· | ···················· | ···················· |
| O | 1922 | hCoV-19/England/CAMC-A3F095/2020\|EPI_ISL_609583\|20 | ···················· | ···················· | ····················· | ···················· | ···················· | ···················· |
| O | 1923 | hCoV-19/England/CAMC-A3F262/2020\|EPI_ISL_609584\|20 | ···················· | ···················· | ····················· | ···················· | ···················· | ···················· |
| O | 1924 | hCoV-19/England/CAMC-A3ECB0/2020\|EPI_ISL_609585\|20 | ···················· | ···················· | ····················· | ···················· | ···················· | ···················· |
| O | 1925 | hCoV-19/England/CAMC-A3F323/2020\|EPI_ISL_609586\|20 | ···················· | ···················· | ····················· | ···················· | ···················· | ···················· |
| O | 1926 | hCoV-19/England/CAMC-A3F226/2020\|EPI_ISL_609587\|20 | ···················· | ·A·················· | ····················· | ···················· | ···················· | ···A················ |
| O | 1927 | hCoV-19/England/CAMC-A3F36F/2020\|EPI_ISL_609588\|20 | ···················· | ···················· | ····················· | ···················· | ···················· | ···················· |
| O | 1928 | hCoV-19/England/CAMC-A3F4C6/2020\|EPI_ISL_609589\|20 | ···················· | ···················· | ····················· | ···················· | ···················· | ···················· |
| O | 1929 | hCoV-19/England/CAMC-A3F1CF/2020\|EPI_ISL_609590\|20 | ···················· | ···················· | ····················· | ···················· | ···················· | ···················· |
| O | 1930 | hCoV-19/England/CAMC-A3EE8D/2020\|EPI_ISL_609591\|20 | ···················· | ···················· | ····················· | ···················· | ···················· | ···················· |
| O | 1931 | hCoV-19/England/CAMC-A3F45D/2020\|EPI_ISL_609592\|20 | ···················· | ···················· | ····················· | ···················· | ···················· | ···················· |
| O | 1932 | hCoV-19/England/CAMC-A3F4B7/2020\|EPI_ISL_609593\|20 | ···················· | ···················· | ····················· | ···················· | ···················· | ···················· |
| O | 1933 | hCoV-19/England/CAMC-A3EC47/2020\|EPI_ISL_609594\|20 | ···················· | ···················· | ····················· | ···················· | ···················· | ···················· |
| O | 1934 | hCoV-19/England/CAMC-A3F7BE/2020\|EPI_ISL_609595\|20 | ···················· | ···················· | ····················· | ···················· | ···················· | ···················· |
| O | 1935 | hCoV-19/England/CAMC-A3E9C8/2020\|EPI_ISL_609596\|20 | ···················· | ···················· | ····················· | ···················· | ···················· | ···················· |
| O | 1936 | hCoV-19/England/CAMC-A3F3C9/2020\|EPI_ISL_609597\|20 | ···················· | ···················· | ····················· | ···················· | ···················· | ···················· |
| O | 1937 | hCoV-19/England/CAMC-A3E597/2020\|EPI_ISL_609598\|20 | ···················· | ···················· | ····················· | ···················· | ···················· | ···················· |
| O | 1938 | hCoV-19/England/CAMC-A3F43F/2020\|EPI_ISL_609599\|20 | ···················· | ···················· | ····················· | ···················· | ···················· | ···················· |
| O | 1939 | hCoV-19/England/ALDP-A85D99/2020\|EPI_ISL_610400\|20 | ···················· | ···················· | ····················· | ···················· | ···················· | ···················· |
| O | 1940 | hCoV-19/England/ALDP-A85F93/2020\|EPI_ISL_610402\|20 | ···················· | ···················· | ····················· | ···················· | ···················· | ···················· |
| O | 1941 | hCoV-19/England/ALDP-A85AFC/2020\|EPI_ISL_610403\|20 | ···················· | ···················· | ····················· | ···················· | ···················· | ···················· |
| O | 1942 | hCoV-19/England/ALDP-A84A1B/2020\|EPI_ISL_610404\|20 | ···················· | ···················· | ····················· | ···················· | ···················· | ···················· |
| O | 1943 | hCoV-19/England/ALDP-A8540D/2020\|EPI_ISL_610405\|20 | ···················· | ···················· | ····················· | ···················· | ···················· | ···················· |
| O | 1944 | hCoV-19/England/ALDP-A84996/2020\|EPI_ISL_610406\|20 | ···················· | ···················· | ····················· | ···················· | ···················· | ···················· |
| O | 1945 | hCoV-19/England/ALDP-A85397/2020\|EPI_ISL_610408\|20 | ···················· | ···················· | ···········Y········· | ···················· | ···················· | ···················· |
| O | 1946 | hCoV-19/England/ALDP-A84B90/2020\|EPI_ISL_610412\|20 | ···················· | ···················· | ····················· | ···················· | ···················· | ···················· |
| O | 1947 | hCoV-19/England/ALDP-A85F0C/2020\|EPI_ISL_610558\|20 | ···················· | ···················· | ····················· | ···················· | ···················· | ···················· |
| O | 1948 | hCoV-19/England/ALDP-A85537/2020\|EPI_ISL_610559\|20 | ···················· | ···················· | ····················· | ···················· | ···················· | ···················· |
| O | 1949 | hCoV-19/England/ALDP-A85A38/2020\|EPI_ISL_610560\|20 | ···················· | ···················· | ····················· | ···················· | ···················· | ···················· |
| O | 1950 | hCoV-19/England/ALDP-A858C5/2020\|EPI_ISL_610561\|20 | ···················· | ···················· | ····················· | ···················· | ···················· | ···················· |
| O | 1951 | hCoV-19/England/ALDP-A84BDC/2020\|EPI_ISL_610562\|20 | ···················· | ···················· | ····················· | ···················· | ···················· | ···················· |
| O | 1952 | hCoV-19/England/ALDP-A85D7B/2020\|EPI_ISL_610563\|20 | ···················· | ···················· | ····················· | ···················· | ···················· | ···················· |
| O | 1953 | hCoV-19/England/ALDP-A857E6/2020\|EPI_ISL_610564\|20 | ···················· | ···················· | ····················· | ···················· | ····T··············· | ···················· |
| O | 1954 | hCoV-19/England/ALDP-A84CF7/2020\|EPI_ISL_610565\|20 | ···················· | ···················· | ····················· | ···················· | ···················· | ···················· |
| O | 1955 | hCoV-19/England/ALDP-A85DC6/2020\|EPI_ISL_610566\|20 | ···················· | ···················· | ····················· | ···················· | ···················· | ···················· |
| O | 1956 | hCoV-19/England/ALDP-A85E69/2020\|EPI_ISL_610567\|20 | ···················· | ···················· | ····················· | ···················· | ···················· | ···················· |
| O | 1957 | hCoV-19/England/ALDP-A85476/2020\|EPI_ISL_610568\|20 | ···················· | ···················· | ····················· | ···················· | ···················· | ···················· |
| O | 1958 | hCoV-19/England/ALDP-A84987/2020\|EPI_ISL_610569\|20 | ···················· | ···················· | ····················· | ···················· | ···················· | ···················· |
| O | 1959 | hCoV-19/England/ALDP-A85379/2020\|EPI_ISL_610570\|20 | ···················· | ···················· | ····················· | ···················· | ···················· | ···················· |
| O | 1960 | hCoV-19/England/ALDP-A859C2/2020\|EPI_ISL_610572\|20 | ···················· | ···················· | ····················· | ···················· | ···················· | ···················· |
| O | 1961 | hCoV-19/England/ALDP-A84A57/2020\|EPI_ISL_610573\|20 | ···················· | ···················· | ····················· | ···················· | ···················· | ···················· |
| O | 1962 | hCoV-19/England/ALDP-A84F94/2020\|EPI_ISL_610575\|20 | ···················· | ···················· | ····················· | ···················· | ···················· | ···················· |
| O | 1963 | hCoV-19/England/ALDP-A85494/2020\|EPI_ISL_610576\|20 | ···················· | ···················· | ····················· | ···················· | ···················· | ···················· |
| O | 1964 | hCoV-19/England/ALDP-A850EB/2020\|EPI_ISL_610578\|20 | ···················· | ···················· | ···········T········· | ···················· | ···················· | ···················· |
| O | 1965 | hCoV-19/England/ALDP-A85081/2020\|EPI_ISL_610579\|20 | ···················· | ···················· | ····················· | ···················· | ···················· | ···················· |
| O | 1966 | hCoV-19/England/ALDP-A84FEF/2020\|EPI_ISL_610580\|20 | ···················· | ···················· | ····················· | ···················· | ···················· | ···················· |
| O | 1967 | hCoV-19/England/ALDP-A85EB4/2020\|EPI_ISL_610581\|20 | ···················· | ·A·················· | ····················· | ···················· | ···················· | ···················· |
| O | 1968 | hCoV-19/England/ALDP-A8493C/2020\|EPI_ISL_610582\|20 | ···················· | ···················· | ····················· | ···················· | ···················· | ···················· |
| O | 1969 | hCoV-19/England/ALDP-A85CAB/2020\|EPI_ISL_610583\|20 | ···················· | ···················· | ····················· | ···················· | ···················· | ···················· |
| O | 1970 | hCoV-19/England/ALDP-A85C41/2020\|EPI_ISL_610584\|20 | ···················· | ···················· | ····················· | ···················· | ···················· | ···················· |
| O | 1971 | hCoV-19/England/ALDP-A85EA5/2020\|EPI_ISL_610585\|20 | ···················· | ···················· | ····················· | ···················· | ···················· | ···················· |
| O | 1972 | hCoV-19/England/ALDP-A84D7C/2020\|EPI_ISL_610586\|20 | ···················· | ···················· | ····················· | ···················· | ···················· | ···················· |
| O | 1973 | hCoV-19/England/ALDP-A85467/2020\|EPI_ISL_610587\|20 | ···················· | ···················· | ····················· | ···················· | ···················· | ···A················ |
| O | 1974 | hCoV-19/England/ALDP-A85F57/2020\|EPI_ISL_610588\|20 | ···················· | ···················· | ····················· | ···················· | ···················· | ···················· |
| O | 1975 | hCoV-19/England/ALDP-A85203/2020\|EPI_ISL_610589\|20 | ···················· | ···················· | ····················· | ···················· | ···················· | ···················· |
| O | 1976 | hCoV-19/England/ALDP-A84D4F/2020\|EPI_ISL_610594\|20 | ···················· | ···················· | ····················· | ···················· | ···················· | ···················· |
| O | 1977 | hCoV-19/England/ALDP-A85FB1/2020\|EPI_ISL_610595\|20 | ···················· | ···················· | ····················· | ···················· | ······T············· | ···················· |
| O | 1978 | hCoV-19/England/ALDP-A85E1E/2020\|EPI_ISL_610596\|20 | ···················· | ···················· | ····················· | ···················· | ···················· | ···················· |
| O | 1979 | hCoV-19/England/ALDP-A85B08/2020\|EPI_ISL_610597\|20 | ···················· | ···················· | ····················· | ···················· | ···················· | ···················· |
| O | 1980 | hCoV-19/England/ALDP-A85DD5/2020\|EPI_ISL_610598\|20 | ···················· | ···················· | ····················· | ···················· | ···················· | ···················· |
| O | 1981 | hCoV-19/England/ALDP-A84E00/2020\|EPI_ISL_610599\|20 | ···················· | ···················· | ····················· | ···················· | ···················· | ···················· |
| O | 1982 | hCoV-19/USA/VA-DCLS-1601/2020\|EPI_ISL_614012\|2020- | ···················· | ···················· | ····················· | ···················· | ···················· | ···················· |
| O | 1983 | hCoV-19/USA/VA-DCLS-1602/2020\|EPI_ISL_614013\|2020- | ···················· | ···················· | ····················· | ···················· | ···················· | ···················· |
| O | 1984 | hCoV-19/USA/VA-DCLS-1604/2020\|EPI_ISL_614014\|2020- | ···················· | ···················· | ····················· | ···················· | ···················· | ···················· |
| O | 1985 | hCoV-19/USA/VA-DCLS-1605/2020\|EPI_ISL_614015\|2020- | ···················· | ·A·················· | ····················· | ···················· | ···················· | ···················· |
| O | 1986 | hCoV-19/USA/VA-DCLS-1606/2020\|EPI_ISL_614016\|2020- | ···················· | ·A·················· | ····················· | ···················· | ···················· | ···················· |
| O | 1987 | hCoV-19/USA/VA-DCLS-1607/2020\|EPI_ISL_614017\|2020- | ···················· | ···················· | ····················· | ···················· | ···················· | ···················· |
| O | 1988 | hCoV-19/USA/VA-DCLS-1609/2020\|EPI_ISL_614018\|2020- | ···················· | ···················· | ····················· | ···················· | ···················· | ···················· |
| O | 1989 | hCoV-19/USA/VA-DCLS-1610/2020\|EPI_ISL_614019\|2020- | ···················· | ···················· | ····················· | ···················· | ···················· | ···················· |
| O | 1990 | hCoV-19/USA/VA-DCLS-1612/2020\|EPI_ISL_614020\|2020- | ···················· | ···················· | ····················· | ···················· | ···················· | ···················· |
| O | 1991 | hCoV-19/USA/VA-DCLS-1613/2020\|EPI_ISL_614021\|2020- | ···················· | ···················· | ····················· | ···················· | ···················· | ···················· |

| | | | | | | | |
|---|---|---|---|---|---|---|---|
| O | 1992 hCoV-19/USA/VA-DCLS-1614/2020\|EPI_ISL_614022\|2020- | ···················· | ···················· | ···················· | ···················· | ···················· | ···················· |
| O | 1993 hCoV-19/USA/VA-DCLS-1615/2020\|EPI_ISL_614023\|2020- | ···················· | ·A·················· | ···················· | ···················· | ···················· | ···················· |
| O | 1994 hCoV-19/USA/VA-DCLS-1617/2020\|EPI_ISL_614024\|2020- | ···················· | ···················· | ···················· | ···················· | ···················· | ···················· |
| O | 1995 hCoV-19/USA/VA-DCLS-1618/2020\|EPI_ISL_614025\|2020- | ···················· | ···················· | ···················· | ···················· | ···················· | ···················· |
| O | 1996 hCoV-19/USA/VA-DCLS-1619/2020\|EPI_ISL_614026\|2020- | ···················· | ···················· | ···················· | ···················· | ···················· | ···················· |
| O | 1997 hCoV-19/USA/VA-DCLS-1620/2020\|EPI_ISL_614027\|2020- | ···················· | ···················· | ···················· | ···················· | ···················· | ···················· |
| O | 1998 hCoV-19/USA/VA-DCLS-1621/2020\|EPI_ISL_614028\|2020- | ···················· | ···················· | ···················· | ···················· | ···················· | ···················· |
| O | 1999 hCoV-19/USA/VA-DCLS-1624/2020\|EPI_ISL_614029\|2020- | ···················· | ···················· | ···················· | ···················· | ···················· | ···················· |
| O | 2000 hCoV-19/USA/VA-DCLS-1625/2020\|EPI_ISL_614030\|2020- | ···················· | ···················· | ···················· | ···················· | ···················· | ···················· |
| O | 2001 hCoV-19/USA/VA-DCLS-1626/2020\|EPI_ISL_614031\|2020- | ···················· | ·A·················· | ···················· | ···················· | ···················· | ···················· |
| O | 2002 hCoV-19/USA/VA-DCLS-1629/2020\|EPI_ISL_614032\|2020- | ···················· | ···················· | ···················· | ···················· | ···················· | ···················· |
| O | 2003 hCoV-19/USA/VA-DCLS-1630/2020\|EPI_ISL_614033\|2020- | ···················· | ···················· | ···················· | ···················· | ···················· | ···················· |
| O | 2004 hCoV-19/USA/VA-DCLS-1631/2020\|EPI_ISL_614034\|2020- | ···················· | ···················· | ···················· | ···················· | ···················· | ···················· |
| O | 2005 hCoV-19/USA/VA-DCLS-1632/2020\|EPI_ISL_614035\|2020- | ···················· | ···················· | ···················· | ···················· | ···················· | ···················· |
| O | 2006 hCoV-19/USA/VA-DCLS-1633/2020\|EPI_ISL_614036\|2020- | ···················· | ···················· | ···················· | ···················· | ···················· | ···················· |
| O | 2007 hCoV-19/USA/VA-DCLS-1634/2020\|EPI_ISL_614037\|2020- | ···················· | ···················· | ···················· | ···················· | ···················· | ···················· |
| O | 2008 hCoV-19/USA/VA-DCLS-1635/2020\|EPI_ISL_614038\|2020- | ···················· | ···················· | ···················· | ···················· | ···················· | ···················· |
| O | 2009 hCoV-19/USA/VA-DCLS-1636/2020\|EPI_ISL_614039\|2020- | ···················· | ···················· | ···················· | ···················· | ···················· | ···················· |
| O | 2010 hCoV-19/USA/VA-DCLS-1637/2020\|EPI_ISL_614040\|2020- | ·T·················· | ···················· | ···················· | ···················· | ···················· | ···················· |
| O | 2011 hCoV-19/USA/VA-DCLS-1638/2020\|EPI_ISL_614041\|2020- | ···················· | ···················· | ···················· | ···················· | ···················· | ···················· |
| O | 2012 hCoV-19/USA/VA-DCLS-1639/2020\|EPI_ISL_614042\|2020- | ···················· | ···················· | ···················· | ···················· | ···················· | ···················· |
| O | 2013 hCoV-19/USA/VA-DCLS-1640/2020\|EPI_ISL_614043\|2020- | ···················· | ···················· | ···················· | ···················· | ···················· | ···················· |
| O | 2014 hCoV-19/USA/VA-DCLS-1642/2020\|EPI_ISL_614044\|2020- | ···················· | ···················· | ···················· | ···················· | ···················· | ···················· |
| O | 2015 hCoV-19/USA/VA-DCLS-1643/2020\|EPI_ISL_614045\|2020- | ···················· | ···················· | ···················· | ···················· | ···················· | ···················· |
| O | 2016 hCoV-19/USA/VA-DCLS-1646/2020\|EPI_ISL_614046\|2020- | ···················· | ···················· | ···················· | ···················· | ···················· | ···················· |
| O | 2017 hCoV-19/USA/VA-DCLS-1647/2020\|EPI_ISL_614047\|2020- | ···················· | ·A·················· | ···················· | ···················· | ···················· | ···················· |
| O | 2018 hCoV-19/USA/VA-DCLS-1847/2020\|EPI_ISL_614048\|2020- | ···················· | ···················· | ···················· | ···················· | ···················· | ···················· |
| O | 2019 hCoV-19/USA/VA-DCLS-1846/2020\|EPI_ISL_614049\|2020- | ···················· | ···················· | ···················· | ···················· | ···················· | ···················· |
| O | 2020 hCoV-19/USA/VA-DCLS-1722/2020\|EPI_ISL_614050\|2020- | ···················· | ···A················ | ···················· | ···················· | ···················· | ···················· |
| O | 2021 hCoV-19/USA/VA-DCLS-1724/2020\|EPI_ISL_614051\|2020- | ···················· | ·A·················· | ···················· | ···················· | ···················· | ···················· |
| O | 2022 hCoV-19/USA/VA-DCLS-1729/2020\|EPI_ISL_614052\|2020- | ···················· | ·A·················· | ···················· | ···················· | ···················· | ···················· |
| O | 2023 hCoV-19/USA/VA-DCLS-1733/2020\|EPI_ISL_614053\|2020- | ···················· | ···A················ | ···················· | ···················· | ···················· | ···················· |
| O | 2024 hCoV-19/USA/VA-DCLS-1735/2020\|EPI_ISL_614054\|2020- | ···················· | ·A·················· | ···················· | ···················· | ···················· | ···················· |
| O | 2025 hCoV-19/USA/VA-DCLS-1803/2020\|EPI_ISL_614055\|2020- | ···················· | ···················· | ···················· | ···················· | ···················· | ···················· |
| O | 2026 hCoV-19/USA/VA-DCLS-1804/2020\|EPI_ISL_614056\|2020- | ···················· | ···················· | ···················· | ···················· | ···················· | ···················· |
| O | 2027 hCoV-19/USA/VA-DCLS-1805/2020\|EPI_ISL_614057\|2020- | ···················· | ·A·················· | ···················· | ···················· | ···················· | ···················· |
| O | 2028 hCoV-19/USA/VA-DCLS-1807/2020\|EPI_ISL_614058\|2020- | ···················· | ···················· | ···················· | ···················· | ···················· | ···················· |
| O | 2029 hCoV-19/USA/VA-DCLS-1808/2020\|EPI_ISL_614059\|2020- | ···················· | ···················· | ···················· | ···················· | ···················· | ···················· |
| O | 2030 hCoV-19/USA/VA-DCLS-1809/2020\|EPI_ISL_614060\|2020- | ···················· | ···················· | ···················· | ···················· | ···················· | ···················· |
| O | 2031 hCoV-19/USA/VA-DCLS-1810/2020\|EPI_ISL_614061\|2020- | ···················· | ···················· | ···················· | ···················· | ···················· | ···················· |
| O | 2032 hCoV-19/USA/VA-DCLS-1811/2020\|EPI_ISL_614062\|2020- | ···················· | ···················· | ···················· | ···················· | ···················· | ···················· |
| O | 2033 hCoV-19/USA/VA-DCLS-1915/2020\|EPI_ISL_614063\|2020- | ···················· | ···················· | ···················· | ···················· | ···················· | ···················· |
| O | 2034 hCoV-19/USA/VA-DCLS-1917/2020\|EPI_ISL_614064\|2020- | ···················· | ···················· | ···················· | ···················· | ···················· | ···················· |
| O | 2035 hCoV-19/USA/VA-DCLS-1918/2020\|EPI_ISL_614065\|2020- | ···················· | ···················· | ···················· | ···················· | ···················· | ···················· |
| O | 2036 hCoV-19/USA/VA-DCLS-1919/2020\|EPI_ISL_614066\|2020- | ···················· | ···················· | ···················· | ···················· | ···················· | ···················· |
| O | 2037 hCoV-19/USA/VA-DCLS-1920/2020\|EPI_ISL_614067\|2020- | ···················· | ···················· | ···················· | ···················· | ···················· | ···················· |
| O | 2038 hCoV-19/USA/VA-DCLS-1921/2020\|EPI_ISL_614068\|2020- | ···················· | ···················· | ···················· | ···················· | ···················· | ···················· |
| O | 2039 hCoV-19/USA/VA-DCLS-1922/2020\|EPI_ISL_614069\|2020- | ···················· | ···················· | ···················· | ···················· | ···················· | ···················· |
| O | 2040 hCoV-19/USA/VA-DCLS-1896/2020\|EPI_ISL_614071\|2020- | ···················· | ···················· | ···················· | ···················· | ···················· | ···················· |
| O | 2041 hCoV-19/USA/VA-DCLS-1897/2020\|EPI_ISL_614072\|2020- | ···················· | ···················· | ···················· | ···················· | ···················· | ···················· |
| O | 2042 hCoV-19/USA/VA-DCLS-1987/2020\|EPI_ISL_614073\|2020- | ···················· | ·A·················· | ···················· | ···················· | ···················· | ···················· |
| O | 2043 hCoV-19/USA/VA-DCLS-1989/2020\|EPI_ISL_614074\|2020- | ···················· | ·A·················· | ···················· | ···················· | ···················· | ···················· |
| O | 2044 hCoV-19/USA/VA-DCLS-1849/2020\|EPI_ISL_614075\|2020- | ···················· | ···················· | ···················· | ···················· | ···················· | ···················· |
| O | 2045 hCoV-19/USA/VA-DCLS-1850/2020\|EPI_ISL_614076\|2020- | ···················· | ···················· | ···················· | ···················· | ···················· | ···················· |
| O | 2046 hCoV-19/USA/VA-DCLS-1853/2020\|EPI_ISL_614077\|2020- | ···················· | ···················· | ···················· | ···················· | ···················· | ···················· |
| O | 2047 hCoV-19/USA/VA-DCLS-1854/2020\|EPI_ISL_614078\|2020- | ···················· | ···················· | ···················· | ···················· | ···················· | ···················· |
| O | 2048 hCoV-19/USA/VA-DCLS-1855/2020\|EPI_ISL_614079\|2020- | ···················· | ···················· | ···················· | ···················· | ···················· | ···················· |
| O | 2049 hCoV-19/India/DL-ILBS-1718M/2020\|EPI_ISL_605805\|20 | ···················· | ···················· | ···················· | ···················· | ···················· | ···················· |
| O | 2050 hCoV-19/India/DL-ILBS-1929M/2020\|EPI_ISL_605806\|20 | ···················· | ···················· | ···················· | ···················· | ···················· | ···················· |
| O | 2051 hCoV-19/India/DL-ILBS-2476T/2020\|EPI_ISL_605807\|20 | ···················· | ···················· | ···················· | ···················· | ···················· | ···················· |
| O | 2052 hCoV-19/India/DL-ILBS-271M/2020\|EPI_ISL_605808\|202 | ···················· | ···················· | ···················· | ···················· | ···················· | ···················· |
| O | 2053 hCoV-19/India/DL-ILBS-299M/2020\|EPI_ISL_605809\|202 | ···················· | ···················· | ···················· | ···················· | ···················· | ···················· |
| O | 2054 hCoV-19/India/DL-ILBS-9062D/2020\|EPI_ISL_605811\|20 | ···················· | ···················· | ···················· | ···················· | ···················· | ···················· |
| O | 2055 hCoV-19/India/DL-ILBS-9063D/2020\|EPI_ISL_605812\|20 | ···················· | ···················· | ···················· | ···················· | ···················· | ···················· |
| O | 2056 hCoV-19/India/DL-ILBS-9304D/2020\|EPI_ISL_605813\|20 | ···················· | ···················· | ···················· | ···················· | ···················· | ···················· |
| O | 2057 hCoV-19/India/DL-ILBS-9309D/2020\|EPI_ISL_605814\|20 | ···················· | ·A·················· | ···················· | ···················· | ···················· | ··A················· |
| O | 2058 hCoV-19/India/DL-ILBS-9318D/2020\|EPI_ISL_605815\|20 | ···················· | ·A·················· | ···················· | ···················· | ···················· | ··A················· |
| O | 2059 hCoV-19/England/ALDP-A6CF1A/2020\|EPI_ISL_608080\|20 | ···················· | ···················· | ···················· | ···················· | ···················· | ···················· |
| O | 2060 hCoV-19/England/ALDP-A6D09E/2020\|EPI_ISL_608081\|20 | ···················· | ···················· | ···················· | ···················· | ···················· | ···················· |
| O | 2061 hCoV-19/England/ALDP-A6CFFC/2020\|EPI_ISL_608083\|20 | ···················· | ·A·················· | ···················· | ···················· | ···················· | ··A················· |
| O | 2062 hCoV-19/England/ALDP-A70010/2020\|EPI_ISL_608084\|20 | ···················· | ···················· | ···················· | ···················· | ···················· | ···················· |
| O | 2063 hCoV-19/England/ALDP-A6FADA/2020\|EPI_ISL_608085\|20 | ···················· | ···················· | ···················· | ···················· | ···················· | ···················· |
| O | 2064 hCoV-19/England/ALDP-A7014A/2020\|EPI_ISL_608086\|20 | ···················· | ···················· | ···················· | ···················· | ···················· | ···················· |
| O | 2065 hCoV-19/England/ALDP-A70915/2020\|EPI_ISL_608087\|20 | ···················· | ···················· | ···················· | ···················· | ···················· | ···················· |
| O | 2066 hCoV-19/England/ALDP-A6D6BA/2020\|EPI_ISL_608100\|20 | ···················· | ···················· | ···················· | ···················· | ···················· | ···················· |
| O | 2067 hCoV-19/England/ALDP-A6D74E/2020\|EPI_ISL_608101\|20 | ···················· | ···················· | ···················· | ···················· | ···················· | ···················· |
| O | 2068 hCoV-19/England/ALDP-A6CB43/2020\|EPI_ISL_608102\|20 | ···················· | ···················· | ···················· | ···················· | ···················· | ···················· |
| O | 2069 hCoV-19/England/ALDP-A6C4EE/2020\|EPI_ISL_608103\|20 | ···················· | ···················· | ···················· | ···················· | ···················· | ···················· |
| O | 2070 hCoV-19/England/ALDP-A6D41A/2020\|EPI_ISL_608104\|20 | ···················· | ···················· | ···················· | ···················· | ···················· | ···················· |
| O | 2071 hCoV-19/England/ALDP-A70274/2020\|EPI_ISL_608105\|20 | ···················· | ·A·················· | ···················· | ···················· | ···················· | ··A················· |

| | | | | | | | |
|---|---|---|---|---|---|---|---|
| O | 2072 hCoV-19/England/ALDP-A6D702/2020\|EPI_ISL_608106\|20 | ···················· | ···················· | ···················· | ···················· | ···················· | ···················· |
| O | 2073 hCoV-19/England/ALDP-A6D474/2020\|EPI_ISL_608107\|20 | ···················· | ···················· | ···················· | ···················· | ···················· | ···················· |
| O | 2074 hCoV-19/England/ALDP-A6C40C/2020\|EPI_ISL_608108\|20 | ···················· | ···················· | ···················· | ···················· | ···················· | ···················· |
| O | 2075 hCoV-19/England/ALDP-A6D0E9/2020\|EPI_ISL_608109\|20 | ···················· | ···················· | ···················· | ···················· | ···················· | ···················· |
| O | 2076 hCoV-19/England/ALDP-A6FECF/2020\|EPI_ISL_608110\|20 | ···················· | ···················· | ···················· | ···················· | ···················· | ···················· |
| O | 2077 hCoV-19/England/ALDP-A70229/2020\|EPI_ISL_608111\|20 | ···················· | ·A·················· | ···················· | ···················· | ···················· | ··A················· |
| O | 2078 hCoV-19/England/ALDP-A70256/2020\|EPI_ISL_608112\|20 | ···················· | ···················· | ···················· | ···················· | ···················· | ···················· |
| O | 2079 hCoV-19/England/ALDP-A6FF71/2020\|EPI_ISL_608113\|20 | ···················· | ···················· | ···················· | ···················· | ···················· | ···················· |
| O | 2080 hCoV-19/England/ALDP-A6FF35/2020\|EPI_ISL_608114\|20 | ···················· | ···················· | ···················· | ···················· | ···················· | ···················· |
| O | 2081 hCoV-19/England/ALDP-A6D3B3/2020\|EPI_ISL_608115\|20 | ···················· | ···················· | ···················· | ···················· | ···················· | ···················· |
| O | 2082 hCoV-19/England/ALDP-A6C615/2020\|EPI_ISL_608116\|20 | ···················· | ···················· | ···················· | ···················· | ···················· | ···················· |
| O | 2083 hCoV-19/England/ALDP-A7020B/2020\|EPI_ISL_608118\|20 | ···················· | ·A·················· | ···················· | ···················· | ···················· | ··A················· |
| O | 2084 hCoV-19/England/ALDP-A6CDE3/2020\|EPI_ISL_608119\|20 | ···················· | ···················· | ···················· | ···················· | ···················· | ···················· |
| O | 2085 hCoV-19/England/ALDP-A70195/2020\|EPI_ISL_608120\|20 | ···················· | ···················· | ···················· | ···················· | ···················· | ···················· |
| O | 2086 hCoV-19/England/ALDP-A6FA9E/2020\|EPI_ISL_608121\|20 | ···················· | ···················· | ···················· | ···················· | ···················· | ···················· |
| O | 2087 hCoV-19/England/ALDP-A6D368/2020\|EPI_ISL_608122\|20 | ···················· | ···················· | ···················· | ···················· | ···················· | ···················· |
| O | 2088 hCoV-19/England/ALDP-A700C5/2020\|EPI_ISL_608123\|20 | ···················· | ···················· | ···················· | ···················· | ···················· | ···················· |
| O | 2089 hCoV-19/England/ALDP-A6D6C9/2020\|EPI_ISL_608124\|20 | ···················· | ···················· | ···················· | ···················· | ···················· | ···················· |
| O | 2090 hCoV-19/England/ALDP-A6CF83/2020\|EPI_ISL_608125\|20 | ···················· | ···················· | ···················· | ···················· | ···················· | ···················· |
| O | 2091 hCoV-19/England/ALDP-A70793/2020\|EPI_ISL_608126\|20 | ···················· | ···················· | ···················· | ···················· | ···················· | ···················· |
| O | 2092 hCoV-19/England/ALDP-A6D6D8/2020\|EPI_ISL_608127\|20 | ···················· | ···················· | ···················· | ···················· | ···················· | ···················· |
| O | 2093 hCoV-19/England/ALDP-A6CF0B/2020\|EPI_ISL_608128\|20 | ···················· | ···················· | ···················· | ···················· | ···················· | ···················· |
| O | 2094 hCoV-19/England/ALDP-A6D18C/2020\|EPI_ISL_608129\|20 | ···················· | ···················· | ···················· | ···················· | ···················· | ···················· |
| O | 2095 hCoV-19/England/ALDP-A6D76C/2020\|EPI_ISL_608130\|20 | ···················· | ···················· | ···················· | ···················· | ···················· | ···················· |
| O | 2096 hCoV-19/England/ALDP-A708BE/2020\|EPI_ISL_608131\|20 | ···················· | ···················· | ···················· | ···················· | ···················· | ···················· |
| O | 2097 hCoV-19/England/ALDP-A6D113/2020\|EPI_ISL_608132\|20 | ···················· | ···················· | ···················· | ···················· | ···················· | ···················· |
| O | 2098 hCoV-19/England/ALDP-A6D27A/2020\|EPI_ISL_608133\|20 | ···················· | ···················· | ···················· | ···················· | ···················· | ···················· |
| O | 2099 hCoV-19/England/ALDP-A6D298/2020\|EPI_ISL_608134\|20 | ···················· | ···················· | ···················· | ···················· | ···················· | ···················· |
| O | 2100 hCoV-19/England/ALDP-A6C484/2020\|EPI_ISL_608135\|20 | ···················· | ···················· | ···················· | ···················· | ···················· | ···················· |
| O | 2101 hCoV-19/England/ALDP-A6C5CD/2020\|EPI_ISL_608136\|20 | ···················· | ···················· | ···················· | ···················· | ···················· | ···················· |
| O | 2102 hCoV-19/England/ALDP-A6C91C/2020\|EPI_ISL_608137\|20 | ···················· | ·A·················· | ···················· | ···················· | ···················· | ··A················· |
| O | 2103 hCoV-19/England/ALDP-A707EE/2020\|EPI_ISL_608138\|20 | ···················· | ···················· | ···················· | ···················· | ···················· | ···················· |
| O | 2104 hCoV-19/England/ALDP-A707A2/2020\|EPI_ISL_608139\|20 | ···················· | ·A·······A·········· | ···················· | ···················· | ···················· | ···················· |
| O | 2105 hCoV-19/England/ALDP-A6CB61/2020\|EPI_ISL_608140\|20 | ···················· | ···················· | ···················· | ···················· | ···················· | ···················· |
| O | 2106 hCoV-19/England/ALDP-A6C3B4/2020\|EPI_ISL_608141\|20 | ···················· | ···················· | ···················· | ···················· | ···················· | ···················· |
| O | 2107 hCoV-19/England/ALDP-A6C466/2020\|EPI_ISL_608143\|20 | ···················· | ···················· | ···················· | ···················· | ···················· | ···················· |
| O | 2108 hCoV-19/England/ALDP-A6C457/2020\|EPI_ISL_608145\|20 | ···················· | ···················· | ···················· | ···················· | ···················· | ···················· |
| O | 2109 hCoV-19/England/ALDP-A6D15F/2020\|EPI_ISL_608146\|20 | ···················· | ·A·················· | ···················· | ···················· | ···················· | ··A················· |
| O | 2110 hCoV-19/England/ALDP-A6D7B7/2020\|EPI_ISL_608148\|20 | ···················· | ···················· | ········R··········· | ···················· | ···················· | ···················· |
| O | 2111 hCoV-19/England/ALDP-A6C642/2020\|EPI_ISL_608150\|20 | ···················· | ···················· | ···················· | ···················· | ···················· | ···················· |
| O | 2112 hCoV-19/England/ALDP-A6CB25/2020\|EPI_ISL_608151\|20 | ···················· | ···················· | ···················· | ···················· | ···················· | ···················· |
| O | 2113 hCoV-19/England/ALDP-A6FF26/2020\|EPI_ISL_608152\|20 | ···················· | ·A·················· | ···················· | ···················· | ···················· | ··A················· |
| O | 2114 hCoV-19/England/ALDP-A6CF56/2020\|EPI_ISL_608153\|20 | ···················· | ···················· | ···················· | ···················· | ···················· | ···················· |
| O | 2115 hCoV-19/England/ALDP-A6C8C4/2020\|EPI_ISL_608154\|20 | ···················· | ···················· | ···················· | ···················· | ···················· | ···················· |
| O | 2116 hCoV-19/England/ALDP-A6C3C3/2020\|EPI_ISL_608155\|20 | ···················· | ···················· | ···················· | ···················· | ···················· | ···················· |
| O | 2117 hCoV-19/England/ALDP-A6CB16/2020\|EPI_ISL_608156\|20 | ···················· | ···················· | ···················· | ···················· | ···················· | ···················· |
| O | 2118 hCoV-19/England/ALDP-A6CF29/2020\|EPI_ISL_608157\|20 | ···················· | ·A·················· | ···················· | ···················· | ···················· | ···················· |
| O | 2119 hCoV-19/England/ALDP-A6CCD7/2020\|EPI_ISL_608159\|20 | ···················· | ···················· | ···················· | ···················· | ···T················ | ···················· |
| O | 2120 hCoV-19/England/ALDP-A6D3E0/2020\|EPI_ISL_608160\|20 | ···················· | ···················· | ···················· | ···················· | ···················· | ···················· |
| O | 2121 hCoV-19/England/ALDP-A6C3E1/2020\|EPI_ISL_608161\|20 | ···················· | ···················· | ···················· | ···················· | ···················· | ···················· |
| O | 2122 hCoV-19/England/ALDP-A70283/2020\|EPI_ISL_608162\|20 | ···················· | ···················· | ···················· | ···················· | ···················· | ···················· |
| O | 2123 hCoV-19/England/ALDP-A6CDC5/2020\|EPI_ISL_608163\|20 | ···················· | ···················· | ···················· | ···················· | ···················· | ···················· |
| O | 2124 hCoV-19/England/ALDP-A6C33C/2020\|EPI_ISL_608164\|20 | ···················· | ···················· | ···················· | ···················· | ···················· | ···················· |
| O | 2125 hCoV-19/England/ALDP-A6FFCC/2020\|EPI_ISL_608165\|20 | ···················· | ···················· | ···················· | ···················· | ···················· | ···················· |
| O | 2126 hCoV-19/England/ALDP-A6C8E2/2020\|EPI_ISL_608166\|20 | ···················· | ···················· | ···················· | ···················· | ···················· | ···················· |
| O | 2127 hCoV-19/England/ALDP-A6D2F2/2020\|EPI_ISL_608167\|20 | ···················· | ···················· | ···················· | ···················· | ···················· | ···················· |
| O | 2128 hCoV-19/England/ALDP-A6D4CF/2020\|EPI_ISL_608168\|20 | ···················· | ···················· | ···················· | ···················· | ···················· | ···················· |
| O | 2129 hCoV-19/England/ALDP-A6D016/2020\|EPI_ISL_608169\|20 | ···················· | ···················· | ···················· | ···················· | ···················· | ···················· |
| O | 2130 hCoV-19/England/ALDP-A6D75D/2020\|EPI_ISL_608170\|20 | ···················· | ···················· | ···················· | ···················· | ···················· | ···················· |
| O | 2131 hCoV-19/England/ALDP-A7011D/2020\|EPI_ISL_608171\|20 | ···················· | ···················· | ···················· | ···················· | ···················· | ···················· |
| O | 2132 hCoV-19/England/ALDP-A6C82E/2020\|EPI_ISL_608172\|20 | ···················· | ·A·················· | ···················· | ···················· | ···················· | ··A················· |
| O | 2133 hCoV-19/England/ALDP-A707FD/2020\|EPI_ISL_608173\|20 | ···················· | ···················· | ···················· | ···················· | ···················· | ···················· |
| O | 2134 hCoV-19/England/ALDP-A6D4ED/2020\|EPI_ISL_608174\|20 | ···················· | ···················· | ···················· | ···················· | ···················· | ···················· |
| O | 2135 hCoV-19/England/ALDP-A6C369/2020\|EPI_ISL_608175\|20 | ···················· | ···················· | ···················· | ···················· | ···················· | ···················· |
| O | 2136 hCoV-19/England/ALDP-A6D720/2020\|EPI_ISL_608176\|20 | ···················· | ···················· | ···················· | ···················· | ···················· | ···················· |
| O | 2137 hCoV-19/England/ALDP-A6CEE0/2020\|EPI_ISL_608177\|20 | ···················· | ···················· | ···················· | ···················· | ···················· | ···················· |
| O | 2138 hCoV-19/England/ALDP-A6CED1/2020\|EPI_ISL_608178\|20 | ···················· | ···················· | ···················· | ···················· | ···················· | ···················· |
| O | 2139 hCoV-19/England/ALDP-A6CCE6/2020\|EPI_ISL_608179\|20 | ···················· | ···················· | ···················· | ···················· | ···················· | ···················· |
| O | 2140 hCoV-19/England/ALDP-A6C3D2/2020\|EPI_ISL_608180\|20 | ···················· | ···················· | ···················· | ···················· | ···················· | ···················· |
| O | 2141 hCoV-19/England/ALDP-A6C4DF/2020\|EPI_ISL_608181\|20 | ···················· | ···················· | ···················· | ···················· | ···················· | ···················· |
| O | 2142 hCoV-19/England/ALDP-A701B3/2020\|EPI_ISL_608182\|20 | ···················· | ···················· | ···················· | ···················· | ···················· | ···················· |
| O | 2143 hCoV-19/England/ALDP-A6C8A6/2020\|EPI_ISL_608183\|20 | ···················· | ···················· | ···················· | ···················· | ···················· | ···················· |
| O | 2144 hCoV-19/England/ALDP-A6D061/2020\|EPI_ISL_608184\|20 | ···················· | ···················· | ···················· | ···················· | ···················· | ···················· |
| O | 2145 hCoV-19/England/ALDP-A6D043/2020\|EPI_ISL_608185\|20 | ···················· | ···················· | ···················· | ···················· | ···················· | ···················· |
| O | 2146 hCoV-19/England/ALDP-A6C69D/2020\|EPI_ISL_608186\|20 | ···················· | ···················· | ···················· | ···················· | ···················· | ···················· |
| O | 2147 hCoV-19/England/ALDP-A6D711/2020\|EPI_ISL_608187\|20 | ···················· | ···················· | ···················· | ···················· | ···················· | ···················· |
| O | 2148 hCoV-19/England/ALDP-A6CB34/2020\|EPI_ISL_608188\|20 | ···················· | ···················· | ···················· | ···················· | ···················· | ···················· |
| O | 2149 hCoV-19/England/ALDP-A6D31D/2020\|EPI_ISL_608190\|20 | ···················· | ···················· | ···················· | ···················· | ···················· | ···················· |
| O | 2150 hCoV-19/England/ALDP-A6CD98/2020\|EPI_ISL_608192\|20 | ···················· | ···················· | ···················· | ···················· | ···················· | ···················· |
| O | 2151 hCoV-19/England/ALDP-A6CCF5/2020\|EPI_ISL_608193\|20 | ···················· | ···················· | ···················· | ···················· | ···················· | ···················· |

O 2152 hCoV-19/England/ALDP-A6D4FC/2020|EPI_ISL_608194|20 ···················· ···················· ···················· ···················· ···················· ····················
O 2153 hCoV-19/England/ALDP-A700A7/2020|EPI_ISL_608195|20 ···················· ···················· ···················· ···················· ···················· ····················
O 2154 hCoV-19/England/ALDP-A6D40B/2020|EPI_ISL_608197|20 ···················· ···················· ···················· ···················· ···················· ····················
O 2155 hCoV-19/England/ALDP-A6D535/2020|EPI_ISL_608198|20 ···················· ···················· ···················· ···················· ···················· ····················
O 2156 hCoV-19/England/ALDP-A6C7C7/2020|EPI_ISL_608199|20 ···················· ···················· ···················· ···················· ···················· ····················
O 2157 hCoV-19/England/CAMC-A413C4/2020|EPI_ISL_609400|20 ···················· ···················· ···················· ···················· ···················· ····················
O 2158 hCoV-19/England/CAMC-A413F1/2020|EPI_ISL_609401|20 ···················· ···················· ···················· ···················· ···················· ····················
O 2159 hCoV-19/England/CAMC-A4129A/2020|EPI_ISL_609402|20 ···················· ···················· ···················· ···················· ···················· ····················
O 2160 hCoV-19/England/CAMC-A4190E/2020|EPI_ISL_609403|20 ···················· ···················· ···················· ···················· ···················· ····················
O 2161 hCoV-19/England/CAMC-A422E4/2020|EPI_ISL_609404|20 ···················· ···················· ···················· ···················· ···················· ····················
O 2162 hCoV-19/England/CAMC-A41889/2020|EPI_ISL_609405|20 ···················· ···················· ···················· ···················· ···················· ····················
O 2163 hCoV-19/England/CAMC-A416CB/2020|EPI_ISL_609408|20 ···················· ···················· ···················· ···················· ···················· ····················
O 2164 hCoV-19/England/CAMC-A41E69/2020|EPI_ISL_609409|20 ···················· ···················· ···················· ···················· ···················· ····················
O 2165 hCoV-19/England/CAMC-A4231E/2020|EPI_ISL_609420|20 ···················· ···················· ···················· ···················· ···················· ····················
O 2166 hCoV-19/England/CAMC-A41BAE/2020|EPI_ISL_609421|20 ···················· ···················· ···················· ···················· ···················· ····················
O 2167 hCoV-19/England/CAMC-A41D20/2020|EPI_ISL_609422|20 ···················· ···················· ···················· ···················· ···················· ····················
O 2168 hCoV-19/England/CAMC-A42017/2020|EPI_ISL_609430|20 ···················· ···················· ···················· ···················· ···················· ····················
O 2169 hCoV-19/England/CAMC-A41B80/2020|EPI_ISL_609431|20 ···················· ···················· ···················· ···················· ···················· ····················
O 2170 hCoV-19/England/CAMC-A4178C/2020|EPI_ISL_609432|20 ···················· ·A·················· ···················· ···················· ···················· ··A·················
O 2171 hCoV-19/England/CAMC-A40E3D/2020|EPI_ISL_609433|20 ···················· ···················· ···················· ···················· ···················· ····················
O 2172 hCoV-19/England/CAMC-A41F1B/2020|EPI_ISL_609434|20 ···················· ·····A·············· ···················· ···················· ···················· ····················
O 2173 hCoV-19/England/CAMC-A41230/2020|EPI_ISL_609435|20 ···················· ···················· ···················· ···················· ···················· ····················
O 2174 hCoV-19/England/CAMC-A40DE5/2020|EPI_ISL_609436|20 ···················· ···················· ···················· ···················· ···················· ····················
O 2175 hCoV-19/England/CAMC-A41115/2020|EPI_ISL_609437|20 ···················· ···················· ···················· ···················· ···················· ····················
O 2176 hCoV-19/England/CAMC-A41160/2020|EPI_ISL_609438|20 ···················· ···················· ···················· ···················· ···················· ····················
O 2177 hCoV-19/England/CAMC-A40E88/2020|EPI_ISL_609439|20 ···················· ···················· ···················· ···················· ···················· ····················
O 2178 hCoV-19/England/CAMC-A410FA/2020|EPI_ISL_609440|20 ···················· ···················· ···················· ···················· ···················· ····················
O 2179 hCoV-19/England/CAMC-A41898/2020|EPI_ISL_609441|20 ···················· ···················· ···················· ···················· ···················· ····················
O 2180 hCoV-19/England/CAMC-A41027/2020|EPI_ISL_609442|20 ···················· ···················· ···················· ···················· ···················· ····················
O 2181 hCoV-19/England/CAMC-A40D7C/2020|EPI_ISL_609443|20 ···················· ···················· ···················· ···················· ···················· ····················
O 2182 hCoV-19/England/CAMC-A422C6/2020|EPI_ISL_609444|20 ···················· ···················· ···················· ···················· ···················· ····················
O 2183 hCoV-19/England/CAMC-A40E1F/2020|EPI_ISL_609445|20 ····G··············· ···················· ···················· ···················· ···················· ····················
O 2184 hCoV-19/England/CAMC-A41F57/2020|EPI_ISL_609446|20 ···················· ···················· ···················· ···················· ···················· ····················
O 2185 hCoV-19/England/CAMC-A40D8B/2020|EPI_ISL_609447|20 ···················· ···················· ···················· ···················· ···················· ····················
O 2186 hCoV-19/England/CAMC-A41C8D/2020|EPI_ISL_609449|20 ···················· ···················· ···················· ···················· ···················· ····················
O 2187 hCoV-19/England/CAMC-A4169E/2020|EPI_ISL_609451|20 ···················· ·A·················· ···················· ···················· ···················· ··A·················
O 2188 hCoV-19/England/CAMC-A41986/2020|EPI_ISL_609452|20 ···················· ···················· ···················· ···················· ···················· ····················
O 2189 hCoV-19/England/CAMC-A41740/2020|EPI_ISL_609453|20 ···················· ···················· ···················· ···················· ···················· ····················
O 2190 hCoV-19/England/CAMC-A41810/2020|EPI_ISL_609454|20 ···················· ···················· ···················· ···················· ···················· ····················
O 2191 hCoV-19/England/CAMC-A41801/2020|EPI_ISL_609455|20 ···················· ···················· ···················· ···················· ···················· ····················
O 2192 hCoV-19/England/CAMC-A416BC/2020|EPI_ISL_609456|20 ···················· ···················· ···················· ···················· ···················· ····················
O 2193 hCoV-19/England/CAMC-A4142B/2020|EPI_ISL_609457|20 ···················· ···················· ···················· ···················· ···················· ····················
O 2194 hCoV-19/England/CAMC-A41D99/2020|EPI_ISL_609458|20 ···················· ···················· ···················· ···················· ···················· ····················
O 2195 hCoV-19/England/CAMC-A4135B/2020|EPI_ISL_609459|20 ···················· ···················· ···················· ···················· ···················· ····················
O 2196 hCoV-19/England/CAMC-A41133/2020|EPI_ISL_609460|20 ···················· ···················· ···················· ···················· ···················· ····················
O 2197 hCoV-19/England/CAMC-A411D9/2020|EPI_ISL_609461|20 ····G··············· ···················· ···················· ···················· ···················· ····················
O 2198 hCoV-19/England/CAMC-A4177D/2020|EPI_ISL_609462|20 ···················· ···················· ···················· ···················· ···················· ····················
O 2199 hCoV-19/England/CAMC-A4127C/2020|EPI_ISL_609464|20 ···················· ···················· ···················· ···················· ···················· ····················
O 2200 hCoV-19/England/CAMC-A420EA/2020|EPI_ISL_609465|20 ···················· ···················· ···················· ···················· ···················· ····················
O 2201 hCoV-19/England/CAMC-A413A6/2020|EPI_ISL_609466|20 ···················· ···················· ···················· ···················· ···················· ····················
O 2202 hCoV-19/England/CAMC-A41388/2020|EPI_ISL_609467|20 ···················· ···················· ···················· ···················· ···················· ····················
O 2203 hCoV-19/England/CAMC-A41C32/2020|EPI_ISL_609468|20 ···················· ···················· ···················· ···················· ···················· ····················
O 2204 hCoV-19/England/CAMC-A4183E/2020|EPI_ISL_609469|20 ···················· ···················· ···················· ···················· ···················· ····················
O 2205 hCoV-19/England/CAMC-A41221/2020|EPI_ISL_609470|20 ···················· ···················· ···················· ···················· ···················· ····················
O 2206 hCoV-19/England/CAMC-A417AA/2020|EPI_ISL_609471|20 ···················· ···················· ···················· ···················· ···················· ····················
O 2207 hCoV-19/England/CAMC-A41968/2020|EPI_ISL_609473|20 ···················· ···················· ···················· ···················· ···················· ····················
O 2208 hCoV-19/England/CAMC-A40F58/2020|EPI_ISL_609474|20 ···················· ···················· ···················· ···················· ···················· ····················
O 2209 hCoV-19/England/CAMC-A40F49/2020|EPI_ISL_609475|20 ···················· ···················· ···················· ···················· ···················· ····················
O 2210 hCoV-19/England/CAMC-A4191D/2020|EPI_ISL_609476|20 ···················· ···················· ···················· ···················· ···················· ····················
O 2211 hCoV-19/England/CAMC-A41731/2020|EPI_ISL_609477|20 ···················· ·R·················· ···················· ···················· ···················· ··A·················
O 2212 hCoV-19/England/CAMC-A42035/2020|EPI_ISL_609478|20 ···················· ···················· ···················· ···················· ···················· ····················
O 2213 hCoV-19/England/CAMC-A41BBD/2020|EPI_ISL_609479|20 ···················· ···················· ···················· ···················· ···················· ····················
O 2214 hCoV-19/England/CAMC-A4186B/2020|EPI_ISL_609480|20 ···················· ···················· ···················· ···················· ···················· ····················
O 2215 hCoV-19/England/CAMC-A4217E/2020|EPI_ISL_609481|20 ···················· ···················· ···················· ···················· ···················· ····················
O 2216 hCoV-19/England/CAMC-A416E9/2020|EPI_ISL_609484|20 ···················· ···················· ···················· ···················· ···················· ····················
O 2217 hCoV-19/England/CAMC-A416F8/2020|EPI_ISL_609485|20 ···················· ···················· ···················· ···················· ···················· ····················
O 2218 hCoV-19/England/CAMC-A41CAB/2020|EPI_ISL_609486|20 ···················· ···················· ···················· ···················· ···················· ····················
O 2219 hCoV-19/Wales/CAMC-A40DB8/2020|EPI_ISL_609487|2020 ···················· ···················· ···················· ···················· ···················· ····················
O 2220 hCoV-19/England/CAMC-A41704/2020|EPI_ISL_609488|20 ···················· ···················· ···················· ···················· ···················· ····················
O 2221 hCoV-19/England/CAMC-A41090/2020|EPI_ISL_609489|20 ···················· ···················· ···················· ···················· ···················· ····················
O 2222 hCoV-19/England/CAMC-A417B9/2020|EPI_ISL_609490|20 ···················· ···················· ···················· ···················· ···················· ····················
O 2223 hCoV-19/England/CAMC-A41B71/2020|EPI_ISL_609491|20 ···················· ···················· ···················· ···················· ···················· ····················
O 2224 hCoV-19/England/CAMC-A40E97/2020|EPI_ISL_609492|20 ···················· ···················· ···················· ···················· ···················· ····················
O 2225 hCoV-19/England/CAMC-A41C7E/2020|EPI_ISL_609493|20 ···················· ···················· ···················· ···················· ···················· ····················
O 2226 hCoV-19/England/CAMC-A419FF/2020|EPI_ISL_609494|20 ···················· ···················· ···················· ···················· ···················· ····················
O 2227 hCoV-19/England/CAMC-A41ED2/2020|EPI_ISL_609495|20 ···················· ···················· ···················· ···················· ···················· ····················
O 2228 hCoV-19/England/CAMC-A41300/2020|EPI_ISL_609496|20 ···················· ···················· ···················· ···················· ···················· ····················
O 2229 hCoV-19/England/CAMC-A40DF4/2020|EPI_ISL_609497|20 ···················· ·A·················· ···················· ···················· ···················· ····················
O 2230 hCoV-19/England/CAMC-A41A56/2020|EPI_ISL_609498|20 ···················· ···················· ···················· ···················· ···················· ····················
O 2231 hCoV-19/England/CAMC-A420BD/2020|EPI_ISL_609499|20 ···················· ···················· ···················· ···················· ···················· ····················

O 2232 hCoV-19/England/ALDP-A85BF9/2020|EPI_ISL_610409|20 ···················· ···················· ···················· ···················· ···················· ····················
O 2233 hCoV-19/England/ALDP-A84C8E/2020|EPI_ISL_610414|20 ···················· ···················· ···················· ···················· ···················· ····················
O 2234 hCoV-19/England/ALDP-A859D1/2020|EPI_ISL_610415|20 ···················· ···················· ···················· ···················· ···················· ····················
O 2235 hCoV-19/England/ALDP-A8533D/2020|EPI_ISL_610417|20 ···················· ···················· ···················· ···················· ···················· ····················
O 2236 hCoV-19/England/ALDP-A84A48/2020|EPI_ISL_610418|20 ···················· ···················· ···················· ···················· ···················· ····················
O 2237 hCoV-19/England/ALDP-A85D8A/2020|EPI_ISL_610419|20 ···················· ·A·················· ···················· ···················· ···················· ··A·················
O 2238 hCoV-19/England/ALDP-A8526D/2020|EPI_ISL_610420|20 ···················· ···················· ···················· ···················· ···················· ····················
O 2239 hCoV-19/England/ALDP-A85E78/2020|EPI_ISL_610421|20 ···················· ···················· ···················· ···················· ···················· ····················
O 2240 hCoV-19/England/ALDP-A850FA/2020|EPI_ISL_610422|20 ···················· ···A················ ···················· ···················· ···················· ····················
O 2241 hCoV-19/England/ALDP-A8541C/2020|EPI_ISL_610423|20 ···················· ···················· ···················· ···················· ···················· ····················
O 2242 hCoV-19/England/ALDP-A84B36/2020|EPI_ISL_610424|20 ···················· ···················· ···················· ···················· ···················· ····················
O 2243 hCoV-19/England/ALDP-A84D21/2020|EPI_ISL_610425|20 ···················· ···················· ···················· ···················· ···················· ····················
O 2244 hCoV-19/England/ALDP-A85C32/2020|EPI_ISL_610426|20 ···················· ···················· ···················· ···················· ···················· ····················
O 2245 hCoV-19/England/ALDP-A85C14/2020|EPI_ISL_610427|20 ···················· ···················· ···················· ···················· ···················· ····················
O 2246 hCoV-19/England/ALDP-A86035/2020|EPI_ISL_610429|20 ···················· ···················· ···················· ···················· ···················· ····················
O 2247 hCoV-19/England/ALDP-A85E3C/2020|EPI_ISL_610431|20 ···················· ···················· ···················· ···················· ···················· ····················
O 2248 hCoV-19/England/ALDP-A84E5B/2020|EPI_ISL_610432|20 ···················· ···················· ···················· ···················· ···················· ····················
O 2249 hCoV-19/England/ALDP-A84A75/2020|EPI_ISL_610433|20 ···················· ···················· ···················· ···················· ···················· ····················
O 2250 hCoV-19/England/ALDP-A85ACF/2020|EPI_ISL_610434|20 ···················· ···················· ···················· ···················· ···················· ····················
O 2251 hCoV-19/England/ALDP-A85036/2020|EPI_ISL_610435|20 ···················· ···················· ···················· ···················· ···················· ····················
O 2252 hCoV-19/England/ALDP-A8490F/2020|EPI_ISL_610436|20 ···················· ···················· ···················· ···················· ···················· ····················
O 2253 hCoV-19/England/ALDP-A858D4/2020|EPI_ISL_610437|20 ···················· ···················· ···················· ···················· ···················· ····················
O 2254 hCoV-19/England/ALDP-A84BFA/2020|EPI_ISL_610438|20 ···················· ···················· ···················· ···················· ···················· ····················
O 2255 hCoV-19/England/ALDP-A849E1/2020|EPI_ISL_610439|20 ···················· ···················· ···················· ···················· ···················· ····················
O 2256 hCoV-19/England/ALDP-A84A2A/2020|EPI_ISL_610440|20 ···················· ···················· ···················· ···················· ···················· ····················
O 2257 hCoV-19/England/ALDP-A84C15/2020|EPI_ISL_610441|20 ···················· ···················· ···················· ···················· ···················· ····················
O 2258 hCoV-19/England/ALDP-A84DE5/2020|EPI_ISL_610442|20 ···················· ···················· ···················· ···················· ···················· ····················
O 2259 hCoV-19/England/ALDP-A85546/2020|EPI_ISL_610443|20 ···················· ···················· ···················· ···················· ···················· ····················
O 2260 hCoV-19/England/ALDP-A855A0/2020|EPI_ISL_610444|20 ···················· ···················· ···················· ···················· ···················· ····················
O 2261 hCoV-19/England/ALDP-A85713/2020|EPI_ISL_610445|20 ···················· ···················· ···················· ···················· ···················· ····················
O 2262 hCoV-19/England/ALDP-A85555/2020|EPI_ISL_610447|20 ···················· ···················· ···················· ···················· ···················· ····················
O 2263 hCoV-19/England/ALDP-A85652/2020|EPI_ISL_610449|20 ···················· ···················· ···················· ···················· ···················· ····················
O 2264 hCoV-19/England/ALDP-A84D30/2020|EPI_ISL_610450|20 ···················· ···················· ···················· ···················· ···················· ····················
O 2265 hCoV-19/England/ALDP-A856E9/2020|EPI_ISL_610452|20 ···················· ···················· ···················· ···················· ···················· ····················
O 2266 hCoV-19/England/ALDP-A85986/2020|EPI_ISL_610453|20 ···················· ···················· ···················· ···················· ···················· ····················
O 2267 hCoV-19/England/ALDP-A85C23/2020|EPI_ISL_610454|20 ···················· ···················· ···················· ···················· ···················· ····················
O 2268 hCoV-19/England/ALDP-A84DC7/2020|EPI_ISL_610455|20 ···················· ···················· ···················· ···················· ···················· ····················
O 2269 hCoV-19/England/ALDP-A84A39/2020|EPI_ISL_610457|20 ···················· ···················· ···················· ···················· ···················· ····················
O 2270 hCoV-19/England/ALDP-A85E87/2020|EPI_ISL_610459|20 ···················· ···················· ···················· ···················· ···················· ····················
O 2271 hCoV-19/England/ALDP-A84BBE/2020|EPI_ISL_610461|20 ···················· ···················· ···················· ···················· ···················· ····················
O 2272 hCoV-19/England/ALDP-A8576E/2020|EPI_ISL_610462|20 ···················· ···················· ···················· ···················· ···················· ····················
O 2273 hCoV-19/England/ALDP-A85E0F/2020|EPI_ISL_610463|20 ···················· ···················· ···················· ···················· ···················· ····················
O 2274 hCoV-19/England/ALDP-A8584D/2020|EPI_ISL_610464|20 ···················· ···················· ···················· ···················· ···················· ····················
O 2275 hCoV-19/England/ALDP-A8591D/2020|EPI_ISL_610465|20 ···················· ···················· ···················· ···················· ···················· ····················
O 2276 hCoV-19/England/ALDP-A85A65/2020|EPI_ISL_610467|20 ···················· ···················· ···················· ···················· ···················· ····················
O 2277 hCoV-19/England/ALDP-A85DE4/2020|EPI_ISL_610469|20 ···················· ···················· ···················· ···················· ···················· ····················
O 2278 hCoV-19/England/ALDP-A85898/2020|EPI_ISL_610470|20 ···················· ···················· ···················· ···················· ···················· ····················
O 2279 hCoV-19/England/ALDP-A853D3/2020|EPI_ISL_610472|20 ···················· ···················· ···················· ···················· ···················· ····················
O 2280 hCoV-19/England/ALDP-A85C9C/2020|EPI_ISL_610473|20 ···················· ···················· ···················· ···················· ···················· ····················
O 2281 hCoV-19/England/ALDP-A85B53/2020|EPI_ISL_610474|20 ···················· ···················· ···················· ···················· ···················· ····················
O 2282 hCoV-19/England/ALDP-A856DA/2020|EPI_ISL_610475|20 ···················· ···················· ···················· ···················· ···················· ····················
O 2283 hCoV-19/England/ALDP-A85C8D/2020|EPI_ISL_610476|20 ···················· ·A·················· ···················· ···················· ···················· ··A·················
O 2284 hCoV-19/England/ALDP-A84E97/2020|EPI_ISL_610477|20 ···················· ···················· ···················· ···················· ···················· ····················
O 2285 hCoV-19/England/ALDP-A854D0/2020|EPI_ISL_610478|20 ···················· ···················· ···················· ···················· ···················· ····················
O 2286 hCoV-19/England/ALDP-A84EA6/2020|EPI_ISL_610479|20 ···················· ···················· ···················· ···················· ···················· ····················
O 2287 hCoV-19/England/ALDP-A84A0C/2020|EPI_ISL_610480|20 ···················· ···················· ···················· ···················· ···················· ····················
O 2288 hCoV-19/England/ALDP-A84AEE/2020|EPI_ISL_610481|20 ···················· ···················· ···················· ···················· ···················· ····················
O 2289 hCoV-19/England/ALDP-A84ED3/2020|EPI_ISL_610482|20 ···················· ···················· ···················· ···················· ···················· ····················
O 2290 hCoV-19/England/ALDP-A856F8/2020|EPI_ISL_610483|20 ···················· ···················· ···················· ···················· ···················· ····················
O 2291 hCoV-19/England/ALDP-A851BB/2020|EPI_ISL_610484|20 ···················· ·A·················· ···················· ···················· ···················· ····················
O 2292 hCoV-19/England/ALDP-A84F58/2020|EPI_ISL_610485|20 ···················· ···················· ···················· ···················· ···················· ····················
O 2293 hCoV-19/England/ALDP-A85731/2020|EPI_ISL_610486|20 ···················· ···················· ···················· ···················· ···················· ····················
O 2294 hCoV-19/England/ALDP-A85BAE/2020|EPI_ISL_610487|20 ···················· ···················· ···················· ···················· ···················· ····················
O 2295 hCoV-19/England/ALDP-A85FDF/2020|EPI_ISL_610489|20 ···················· ···················· ···················· ···················· ···················· ····················
O 2296 hCoV-19/England/ALDP-A84B09/2020|EPI_ISL_610490|20 ···················· ···················· ···················· ···················· ···················· ····················
O 2297 hCoV-19/England/ALDP-A84EC4/2020|EPI_ISL_610491|20 ···················· ···················· ···················· ···················· ···················· ····················
O 2298 hCoV-19/England/ALDP-A85607/2020|EPI_ISL_610492|20 ···················· ···················· ···················· ···················· ···················· ····················
O 2299 hCoV-19/England/ALDP-A85740/2020|EPI_ISL_610493|20 ···················· ···················· ···················· ···················· ···················· ····················
O 2300 hCoV-19/England/ALDP-A85300/2020|EPI_ISL_610495|20 ···················· ···················· ···················· ···················· ···················· ····················
O 2301 hCoV-19/England/ALDP-A84D12/2020|EPI_ISL_610496|20 ···················· ···················· ···················· ···················· ···················· ····················
O 2302 hCoV-19/England/ALDP-A85B17/2020|EPI_ISL_610497|20 ···················· ···················· ···················· ···················· ···················· ····················
O 2303 hCoV-19/England/ALDP-A84B27/2020|EPI_ISL_610498|20 ···················· ···················· ···················· ···················· ···················· ····················
O 2304 hCoV-19/England/ALDP-A84B81/2020|EPI_ISL_610499|20 ···················· ···················· ···················· ···················· ···················· ····················
O 2305 hCoV-19/India/DL-ILBS-282M/2020|EPI_ISL_605800|202 ···················· ···················· ···················· ···················· ···················· ····················
O 2306 hCoV-19/India/DL-ILBS-2873T/2020|EPI_ISL_605801|20 ···················· ·A·················· ···················· ···················· ···················· ··A·················
O 2307 hCoV-19/India/DL-ILBS-9070D/2020|EPI_ISL_605802|20 ···················· ···················· ···················· ···················· ···················· ····················
O 2308 hCoV-19/India/DL-ILBS-9071D/2020|EPI_ISL_605803|20 ···················· ···················· ···················· ···················· ···················· ····················
O 2309 hCoV-19/India/DL-ILBS-9306T/2020|EPI_ISL_605804|20 ···················· ···················· ···················· ···················· ···················· ····················
O 2310 hCoV-19/India/DL-ILBS-4871T/2020|EPI_ISL_605810|20 ···················· ·A·················· ···················· ···················· ···················· ··A·················
O 2311 hCoV-19/India/DL-ILBS-9337D/2020|EPI_ISL_605816|20 ···················· ·A·················· ···················· ···················· ···················· ··A·················

| | | | | | | | |
|---|---|---|---|---|---|---|---|
| O | 2312 hCoV-19/Singapore/1108/2020\|EPI_ISL_605817\|2020-10 | ···················· | ···················· | ···················· | ···················· | ···················· | ···················· |
| O | 2313 hCoV-19/Singapore/1109/2020\|EPI_ISL_605818\|2020-10 | ···················· | ···················· | ···················· | ···················· | ···················· | ···················· |
| O | 2314 hCoV-19/Singapore/1110/2020\|EPI_ISL_605819\|2020-10 | ···················· | ···················· | ···················· | ···················· | ···················· | ···················· |
| O | 2315 hCoV-19/Singapore/1111/2020\|EPI_ISL_605820\|2020-10 | ···················· | ···················· | ···················· | ···················· | ···················· | ···················· |
| O | 2316 hCoV-19/Singapore/1112/2020\|EPI_ISL_605821\|2020-10 | ···················· | ···················· | ···················· | ···················· | ···················· | ···················· |
| O | 2317 hCoV-19/Singapore/1113/2020\|EPI_ISL_605822\|2020-10 | ···················· | ···················· | ···················· | ···················· | ···················· | ···················· |
| O | 2318 hCoV-19/Singapore/1114/2020\|EPI_ISL_605823\|2020-10 | ···················· | ···················· | ···················· | ···················· | ···················· | ···················· |
| O | 2319 hCoV-19/Singapore/1115/2020\|EPI_ISL_605824\|2020-10 | ···················· | ···················· | ···················· | ···················· | ···················· | ··A················· |
| O | 2320 hCoV-19/Australia/WA348/2020\|EPI_ISL_605825\|2020-1 | ···················· | ···················· | ···················· | ···················· | ···················· | ···················· |
| O | 2321 hCoV-19/Australia/WA349/2020\|EPI_ISL_605826\|2020-1 | ···················· | ···················· | ···················· | ···················· | ··C················· | ···················· |
| O | 2322 hCoV-19/Australia/WA350/2020\|EPI_ISL_605827\|2020-0 | ···················· | ···················· | ···················· | ···················· | ···················· | ···················· |
| O | 2323 hCoV-19/Australia/WA351/2020\|EPI_ISL_605828\|2020-1 | ···················· | ···················· | ···················· | ···················· | ··C················· | ···················· |
| O | 2324 hCoV-19/Australia/WA352/2020\|EPI_ISL_605829\|2020-1 | ···················· | ···················· | ···················· | ···················· | ··C················· | ···················· |
| O | 2325 hCoV-19/Australia/WA353/2020\|EPI_ISL_605830\|2020-1 | ···················· | ···················· | ···················· | ···················· | ··C················· | ···················· |
| O | 2326 hCoV-19/Australia/WA354/2020\|EPI_ISL_605831\|2020-1 | ···················· | ·A·················· | ···················· | ···················· | ···················· | ··A················· |
| O | 2327 hCoV-19/Australia/WA355/2020\|EPI_ISL_605832\|2020-1 | ···················· | ···················· | ···················· | ···················· | ··C················· | ···················· |
| O | 2328 hCoV-19/Australia/WA356/2020\|EPI_ISL_605833\|2020-0 | ···················· | ···················· | ···················· | ···················· | ···················· | ···················· |
| O | 2329 hCoV-19/Australia/WA357/2020\|EPI_ISL_605834\|2020-1 | ···················· | ···················· | ···················· | ···················· | ··C················· | ···················· |
| O | 2330 hCoV-19/Australia/WA358/2020\|EPI_ISL_605835\|2020-1 | ···················· | ···················· | ···················· | ···················· | ··C················· | ···················· |
| O | 2331 hCoV-19/Australia/WA359/2020\|EPI_ISL_605836\|2020-1 | ···················· | ···················· | ···················· | ···················· | ···················· | ···················· |
| O | 2332 hCoV-19/England/ALDP-A6CF74/2020\|EPI_ISL_608040\|20 | ···················· | ···················· | ···················· | ···················· | ···················· | ···················· |
| O | 2333 hCoV-19/England/ALDP-A6C660/2020\|EPI_ISL_608041\|20 | ···················· | ···················· | ···················· | ···················· | ···················· | ···················· |
| O | 2334 hCoV-19/England/ALDP-A6C493/2020\|EPI_ISL_608042\|20 | ···················· | ···················· | ···················· | ···················· | ···················· | ···················· |
| O | 2335 hCoV-19/England/ALDP-A6D6F6/2020\|EPI_ISL_608043\|20 | ···················· | ···················· | ···················· | ···················· | ···················· | ···················· |
| O | 2336 hCoV-19/England/ALDP-A6D85A/2020\|EPI_ISL_608044\|20 | ···················· | ···················· | ···················· | ···················· | ···················· | ···················· |
| O | 2337 hCoV-19/England/ALDP-A6CCC8/2020\|EPI_ISL_608045\|20 | ···················· | ···················· | ···················· | ···················· | ···················· | ···················· |
| O | 2338 hCoV-19/England/ALDP-A6CE4A/2020\|EPI_ISL_608047\|20 | ···················· | ···················· | ···················· | ···················· | ···················· | ···················· |
| O | 2339 hCoV-19/England/ALDP-A6C888/2020\|EPI_ISL_608048\|20 | ···················· | ···················· | ···················· | ···················· | ···················· | ···················· |
| O | 2340 hCoV-19/England/ALDP-A6C897/2020\|EPI_ISL_608049\|20 | ···················· | ···················· | ···················· | ···················· | ···················· | ···················· |
| O | 2341 hCoV-19/England/ALDP-A6C8D3/2020\|EPI_ISL_608051\|20 | ···················· | ···················· | ···················· | ···················· | ···················· | ···················· |
| O | 2342 hCoV-19/England/ALDP-A6D81E/2020\|EPI_ISL_608052\|20 | ···················· | ···················· | ···················· | ···················· | ···················· | ···················· |
| O | 2343 hCoV-19/England/ALDP-A6D1C8/2020\|EPI_ISL_608053\|20 | ···················· | ···················· | ···················· | ···················· | ···················· | ···················· |
| O | 2344 hCoV-19/England/ALDP-A6D104/2020\|EPI_ISL_608055\|20 | ···················· | ···················· | ···················· | ···················· | ···················· | ···················· |
| O | 2345 hCoV-19/England/ALDP-A6C6D9/2020\|EPI_ISL_608056\|20 | ···················· | ···················· | ···················· | ···················· | ···················· | ···················· |
| O | 2346 hCoV-19/England/ALDP-A7003E/2020\|EPI_ISL_608057\|20 | ···················· | ···················· | ···················· | ···················· | ···················· | ···················· |
| O | 2347 hCoV-19/England/ALDP-A701FF/2020\|EPI_ISL_608058\|20 | ···················· | ···················· | ···················· | ···················· | ···················· | ···················· |
| O | 2348 hCoV-19/England/ALDP-A6C6F7/2020\|EPI_ISL_608059\|20 | ···················· | ···················· | ···················· | ···················· | ···················· | ···················· |
| O | 2349 hCoV-19/England/ALDP-A708FA/2020\|EPI_ISL_608060\|20 | ···················· | ···················· | ···················· | ···················· | ···················· | ···················· |
| O | 2350 hCoV-19/England/ALDP-A6FE1A/2020\|EPI_ISL_608061\|20 | ···················· | ···················· | ···················· | ···················· | ···················· | ···················· |
| O | 2351 hCoV-19/England/ALDP-A6CDB6/2020\|EPI_ISL_608062\|20 | ···················· | ···················· | ···················· | ···················· | ···················· | ···················· |
| O | 2352 hCoV-19/England/ALDP-A6D4DE/2020\|EPI_ISL_608063\|20 | ···················· | ···················· | ···················· | ···················· | ···················· | ···················· |
| O | 2353 hCoV-19/England/ALDP-A700F2/2020\|EPI_ISL_608064\|20 | ···················· | ···················· | ···················· | ···················· | ···················· | ···················· |
| O | 2354 hCoV-19/England/ALDP-A6FAF8/2020\|EPI_ISL_608065\|20 | ···················· | ···················· | ···················· | ···················· | ···················· | ···················· |
| O | 2355 hCoV-19/England/ALDP-A6FB13/2020\|EPI_ISL_608066\|20 | ···················· | ···················· | ···················· | ···················· | ···················· | ···················· |
| O | 2356 hCoV-19/England/ALDP-A701D1/2020\|EPI_ISL_608067\|20 | ···················· | ···················· | ···················· | ···················· | ···················· | ···················· |
| O | 2357 hCoV-19/England/ALDP-A70098/2020\|EPI_ISL_608068\|20 | ···················· | ···················· | ···················· | ···················· | ···················· | ···················· |
| O | 2358 hCoV-19/England/ALDP-A6C624/2020\|EPI_ISL_608070\|20 | ···················· | ···················· | ···················· | ···················· | ···················· | ···················· |
| O | 2359 hCoV-19/England/ALDP-A6D140/2020\|EPI_ISL_608071\|20 | ···················· | ···················· | ···················· | ···················· | ···················· | ···················· |
| O | 2360 hCoV-19/England/ALDP-A6FAE9/2020\|EPI_ISL_608072\|20 | ···················· | ···················· | ···················· | ···················· | ···················· | ···················· |
| O | 2361 hCoV-19/England/ALDP-A6C5BE/2020\|EPI_ISL_608073\|20 | ···················· | ···················· | ···················· | ···················· | ···················· | ···················· |
| O | 2362 hCoV-19/England/ALDP-A6D359/2020\|EPI_ISL_608074\|20 | ···················· | ···················· | ···················· | ···················· | ···················· | ···················· |
| O | 2363 hCoV-19/England/ALDP-A6FE29/2020\|EPI_ISL_608075\|20 | ···················· | ···················· | ···················· | ···················· | ···················· | ···················· |
| O | 2364 hCoV-19/England/ALDP-A6D0DA/2020\|EPI_ISL_608076\|20 | ···················· | ···················· | ···················· | ···················· | ···················· | ···················· |
| O | 2365 hCoV-19/England/ALDP-A6C8F1/2020\|EPI_ISL_608077\|20 | ···················· | ···················· | ···················· | ···················· | ···················· | ···················· |
| O | 2366 hCoV-19/England/ALDP-A6D070/2020\|EPI_ISL_608079\|20 | ···················· | ···················· | ···················· | ···················· | ···················· | ···················· |
| O | 2367 hCoV-19/England/ALDP-A6CE1D/2020\|EPI_ISL_608088\|20 | ···················· | ···················· | ···················· | ···················· | ···················· | ···················· |
| O | 2368 hCoV-19/England/ALDP-A6CE3B/2020\|EPI_ISL_608090\|20 | ···················· | ···················· | ···················· | ···················· | ···················· | ···················· |
| O | 2369 hCoV-19/England/ALDP-A6CEFF/2020\|EPI_ISL_608091\|20 | ···················· | ·A·················· | ···················· | ···················· | ···················· | ···················· |
| O | 2370 hCoV-19/England/ALDP-A7006B/2020\|EPI_ISL_608092\|20 | ···················· | ···················· | ···················· | ···················· | ···················· | ···················· |
| O | 2371 hCoV-19/England/ALDP-A6D289/2020\|EPI_ISL_608093\|20 | ···················· | ···················· | ···················· | ···················· | ···················· | ···················· |
| O | 2372 hCoV-19/England/ALDP-A700D4/2020\|EPI_ISL_608094\|20 | ···················· | ···················· | ···················· | ···················· | ···················· | ···················· |
| O | 2373 hCoV-19/England/ALDP-A6CB07/2020\|EPI_ISL_608096\|20 | ···················· | ···················· | ···················· | ···················· | ···················· | ···················· |
| O | 2374 hCoV-19/England/ALDP-A6D7E4/2020\|EPI_ISL_608098\|20 | ···················· | ···················· | ···················· | ···················· | ···················· | ···················· |
| O | 2375 hCoV-19/England/ALDP-A6D465/2020\|EPI_ISL_608099\|20 | ···················· | ···················· | ···················· | ···················· | ···················· | ···················· |
| O | 2376 hCoV-19/Wales/MILK-A44C5D/2020\|EPI_ISL_609300\|2020 | ···················· | ···················· | ···················· | ···················· | ···················· | ···················· |
| O | 2377 hCoV-19/Wales/MILK-A4675A/2020\|EPI_ISL_609301\|2020 | ···················· | ···················· | ···················· | ···················· | ···················· | ···················· |
| O | 2378 hCoV-19/Wales/MILK-A45C01/2020\|EPI_ISL_609303\|2020 | ···················· | ···················· | ···················· | ···················· | ···················· | ···················· |
| O | 2379 hCoV-19/Wales/MILK-A46848/2020\|EPI_ISL_609304\|2020 | ···················· | ···················· | ···················· | ···················· | ···················· | ···················· |
| O | 2380 hCoV-19/Wales/MILK-A46408/2020\|EPI_ISL_609305\|2020 | ···················· | ···················· | ···················· | ···················· | ···················· | ···················· |
| O | 2381 hCoV-19/Wales/MILK-A46839/2020\|EPI_ISL_609306\|2020 | ···················· | ···················· | ···················· | ···················· | ···················· | ···················· |
| O | 2382 hCoV-19/Wales/MILK-A44D2D/2020\|EPI_ISL_609307\|2020 | ···················· | ···················· | ···················· | ···················· | ···················· | ···················· |
| O | 2383 hCoV-19/England/CAMC-A41494/2020\|EPI_ISL_609308\|20 | ···················· | ···················· | ···················· | ···················· | ···················· | ···················· |
| O | 2384 hCoV-19/England/CAMC-A40ED3/2020\|EPI_ISL_609309\|20 | ···················· | ···················· | ···················· | ···················· | ···················· | ···················· |
| O | 2385 hCoV-19/England/CAMC-A40F1C/2020\|EPI_ISL_609310\|20 | ···················· | ···················· | ···················· | ···················· | ···················· | ···················· |
| O | 2386 hCoV-19/England/CAMC-A40EE2/2020\|EPI_ISL_609311\|20 | ···················· | ···················· | ···················· | ···················· | ···················· | ···················· |
| O | 2387 hCoV-19/England/CAMC-A40F94/2020\|EPI_ISL_609312\|20 | ···················· | ·A·················· | ···················· | ···················· | ···················· | ···················· |
| O | 2388 hCoV-19/England/CAMC-A41081/2020\|EPI_ISL_609313\|20 | ···················· | ···················· | ···················· | ···················· | ···················· | ···················· |
| O | 2389 hCoV-19/England/CAMC-A42026/2020\|EPI_ISL_609314\|20 | ···················· | ···················· | ···················· | ···················· | ···················· | ···················· |
| O | 2390 hCoV-19/England/CAMC-A41EA5/2020\|EPI_ISL_609316\|20 | ···················· | ···················· | ···················· | ···················· | ···················· | ···················· |
| O | 2391 hCoV-19/England/CAMC-A41151/2020\|EPI_ISL_609317\|20 | ···················· | ···················· | ···················· | ···················· | ···················· | ···················· |

| | | | | | | | | |
|---|---|---|---|---|---|---|---|---|
| O | 2392 | hCoV-19/England/CAMC-A42062/2020\|EPI_ISL_609318\|20 | ···················· | ···················· | ····················· | ···················· | ···················· | ···················· |
| O | 2393 | hCoV-19/England/CAMC-A41DF3/2020\|EPI_ISL_609319\|20 | ···················· | ·A·················· | ····················· | ···················· | ···················· | ···················· |
| O | 2394 | hCoV-19/England/CAMC-A41D5D/2020\|EPI_ISL_609320\|20 | ···················· | ···················· | ····················· | ···················· | ···················· | ···················· |
| O | 2395 | hCoV-19/England/CAMC-A411E8/2020\|EPI_ISL_609321\|20 | ···················· | ·A·······A·········· | ····················· | ···················· | ···················· | ···················· |
| O | 2396 | hCoV-19/England/CAMC-A41DD5/2020\|EPI_ISL_609322\|20 | ···················· | ···················· | ····················· | ···················· | ···················· | ···················· |
| O | 2397 | hCoV-19/England/CAMC-A41C41/2020\|EPI_ISL_609323\|20 | ···················· | ·A·················· | ····················· | ···················· | ···················· | ···A················ |
| O | 2398 | hCoV-19/England/CAMC-A41B35/2020\|EPI_ISL_609324\|20 | ···················· | ···················· | ····················· | ···················· | ···················· | ···················· |
| O | 2399 | hCoV-19/England/CAMC-A41995/2020\|EPI_ISL_609326\|20 | ···················· | ···················· | ····················· | ···················· | ···················· | ···················· |
| O | 2400 | hCoV-19/England/CAMC-A4209F/2020\|EPI_ISL_609327\|20 | ···················· | ···················· | ····················· | ···················· | ···················· | ···················· |
| O | 2401 | hCoV-19/England/CAMC-A418E3/2020\|EPI_ISL_609328\|20 | ···················· | ···················· | ····················· | ···················· | ···················· | ···················· |
| O | 2402 | hCoV-19/England/CAMC-A41397/2020\|EPI_ISL_609329\|20 | ···················· | ···················· | ····················· | ···················· | ···················· | ···················· |
| O | 2403 | hCoV-19/England/CAMC-A41EC3/2020\|EPI_ISL_609330\|20 | ···················· | ···················· | ····················· | ···················· | ···················· | ···················· |
| O | 2404 | hCoV-19/England/CAMC-A41FA2/2020\|EPI_ISL_609370\|20 | ···················· | ···················· | ····················· | ···················· | ···················· | ···················· |
| O | 2405 | hCoV-19/England/CAMC-A40FA3/2020\|EPI_ISL_609371\|20 | ···················· | ···················· | ····················· | ···················· | ···················· | ···················· |
| O | 2406 | hCoV-19/England/CAMC-A40D6D/2020\|EPI_ISL_609372\|20 | ···················· | ···················· | ····················· | ···················· | ···················· | ···················· |
| O | 2407 | hCoV-19/England/CAMC-A41DA8/2020\|EPI_ISL_609373\|20 | ···················· | ···················· | ····················· | ···················· | ···················· | ···················· |
| O | 2408 | hCoV-19/England/CAMC-A41D6C/2020\|EPI_ISL_609374\|20 | ···················· | ···················· | ····················· | ···················· | ···················· | ···················· |
| O | 2409 | hCoV-19/England/CAMC-A413E2/2020\|EPI_ISL_609375\|20 | ···················· | ···················· | ····················· | ···················· | ···················· | ···················· |
| O | 2410 | hCoV-19/England/CAMC-A41379/2020\|EPI_ISL_609376\|20 | ···················· | ···················· | ····················· | ···················· | ···················· | ···················· |
| O | 2411 | hCoV-19/England/CAMC-A40F2B/2020\|EPI_ISL_609377\|20 | ···················· | ···················· | ····················· | ···················· | ···················· | ···················· |
| O | 2412 | hCoV-19/England/CAMC-A40F85/2020\|EPI_ISL_609379\|20 | ···················· | ···················· | ····················· | ···················· | ···················· | ···················· |
| O | 2413 | hCoV-19/England/CAMC-A42044/2020\|EPI_ISL_609380\|20 | ···················· | ···················· | ····················· | ···················· | ···················· | ···················· |
| O | 2414 | hCoV-19/England/CAMC-A41F39/2020\|EPI_ISL_609382\|20 | ···················· | ·A·················· | ····················· | ···················· | ···················· | ···A················ |
| O | 2415 | hCoV-19/England/CAMC-A41072/2020\|EPI_ISL_609383\|20 | ···················· | ···················· | ····················· | ···················· | ···················· | ···················· |
| O | 2416 | hCoV-19/England/CAMC-A40DA9/2020\|EPI_ISL_609384\|20 | ···················· | ···················· | ····················· | ···················· | ···················· | ···················· |
| O | 2417 | hCoV-19/England/CAMC-A41F66/2020\|EPI_ISL_609385\|20 | ···················· | ···················· | ····················· | ···················· | ···················· | ···················· |
| O | 2418 | hCoV-19/England/CAMC-A4143A/2020\|EPI_ISL_609386\|20 | ···················· | ···················· | ····················· | ···················· | ···················· | ···················· |
| O | 2419 | hCoV-19/England/CAMC-A41BCC/2020\|EPI_ISL_609387\|20 | ···················· | ·A·················· | ····················· | ···················· | ···················· | ···A················ |
| O | 2420 | hCoV-19/England/CAMC-A41B9F/2020\|EPI_ISL_609388\|20 | ···················· | ···················· | ····················· | ···················· | ···················· | ···················· |
| O | 2421 | hCoV-19/England/CAMC-A41CE7/2020\|EPI_ISL_609389\|20 | ···················· | ···················· | ····················· | ···················· | ···················· | ···················· |
| O | 2422 | hCoV-19/England/CAMC-A41F2A/2020\|EPI_ISL_609390\|20 | ···················· | ···················· | ····················· | ···················· | ···················· | ···················· |
| O | 2423 | hCoV-19/England/CAMC-A41BF9/2020\|EPI_ISL_609391\|20 | ···················· | ···················· | ····················· | ···················· | ···················· | ···················· |
| O | 2424 | hCoV-19/England/CAMC-A41959/2020\|EPI_ISL_609392\|20 | ···················· | ·A·················· | ····················· | ···················· | ···················· | ···A················ |
| O | 2425 | hCoV-19/England/CAMC-A421D8/2020\|EPI_ISL_609394\|20 | ···················· | ···················· | ····················· | ···················· | ···················· | ···················· |
| O | 2426 | hCoV-19/England/CAMC-A40D9A/2020\|EPI_ISL_609395\|20 | ···················· | ···················· | ····················· | ···················· | ···················· | ···················· |
| O | 2427 | hCoV-19/England/CAMC-A4193B/2020\|EPI_ISL_609396\|20 | ···················· | ···················· | ····················· | ···················· | ···················· | ···················· |
| O | 2428 | hCoV-19/England/CAMC-A411AC/2020\|EPI_ISL_609397\|20 | ···················· | ···················· | ····················· | ···················· | ···················· | ···················· |
| O | 2429 | hCoV-19/England/CAMC-A417F5/2020\|EPI_ISL_609398\|20 | ···················· | ···················· | ····················· | ···················· | ···················· | ···················· |
| O | 2430 | hCoV-19/England/CAMC-A41142/2020\|EPI_ISL_609399\|20 | ···················· | ···················· | ····················· | ···················· | ···················· | ···················· |
| O | 2431 | hCoV-19/England/ALDP-A853B5/2020\|EPI_ISL_610300\|20 | ···················· | ···················· | ····················· | ···················· | ···················· | ···················· |
| O | 2432 | hCoV-19/England/ALDP-A84CAC/2020\|EPI_ISL_610301\|20 | ···················· | ···················· | ····················· | ···················· | ···················· | ···················· |
| O | 2433 | hCoV-19/England/ALDP-A8495A/2020\|EPI_ISL_610302\|20 | ···················· | ···················· | ····················· | ···················· | ···················· | ···················· |
| O | 2434 | hCoV-19/England/ALDP-A8593B/2020\|EPI_ISL_610303\|20 | ···················· | ···················· | ····················· | ···················· | ···················· | ···················· |
| O | 2435 | hCoV-19/England/ALDP-A85C6F/2020\|EPI_ISL_610304\|20 | ···················· | ···················· | ····················· | ···················· | ···················· | ···················· |
| O | 2436 | hCoV-19/England/ALDP-A855BF/2020\|EPI_ISL_610306\|20 | ···················· | ···················· | ····················· | ···················· | ···················· | ···················· |
| O | 2437 | hCoV-19/England/ALDP-A8543A/2020\|EPI_ISL_610308\|20 | ···················· | ·········T·········· | ····················· | ···················· | ···················· | ···················· |
| O | 2438 | hCoV-19/England/ALDP-A8568F/2020\|EPI_ISL_610310\|20 | ···················· | ···················· | ····················· | ···················· | ···················· | ···················· |
| O | 2439 | hCoV-19/England/ALDP-A852B8/2020\|EPI_ISL_610311\|20 | ···················· | ···················· | ····················· | ···················· | ···················· | ···················· |
| O | 2440 | hCoV-19/England/ALDP-A8494B/2020\|EPI_ISL_610313\|20 | ···················· | ···················· | ····················· | ···················· | ···················· | ···················· |
| O | 2441 | hCoV-19/England/ALDP-A85564/2020\|EPI_ISL_610314\|20 | ···················· | ···················· | ····················· | ···················· | ···················· | ···················· |
| O | 2442 | hCoV-19/England/ALDP-A85FFD/2020\|EPI_ISL_610320\|20 | ···················· | ···················· | ····················· | ···················· | ···················· | ···················· |
| O | 2443 | hCoV-19/England/ALDP-A85968/2020\|EPI_ISL_610321\|20 | ···················· | ···················· | ····················· | ···················· | ···················· | ···················· |
| O | 2444 | hCoV-19/England/ALDP-A850AF/2020\|EPI_ISL_610360\|20 | ···················· | ···················· | ····················· | ···················· | ···················· | ···················· |
| O | 2445 | hCoV-19/England/ALDP-A8569E/2020\|EPI_ISL_610361\|20 | ···················· | ···················· | ····················· | ···················· | ···················· | ···A················ |
| O | 2446 | hCoV-19/England/ALDP-A85995/2020\|EPI_ISL_610363\|20 | ···················· | ···················· | ····················· | ···················· | ···················· | ···················· |
| O | 2447 | hCoV-19/England/ALDP-A85485/2020\|EPI_ISL_610364\|20 | ···················· | ···················· | ····················· | ···················· | ···················· | ···················· |
| O | 2448 | hCoV-19/England/ALDP-A84DB8/2020\|EPI_ISL_610365\|20 | ···················· | ···················· | ····················· | ···················· | ···················· | ···················· |
| O | 2449 | hCoV-19/England/ALDP-A84DD6/2020\|EPI_ISL_610366\|20 | ···················· | ···················· | ····················· | ···················· | ···················· | ···················· |
| O | 2450 | hCoV-19/England/ALDP-A85AED/2020\|EPI_ISL_610367\|20 | ···················· | ···················· | ····················· | ···················· | ···················· | ···················· |
| O | 2451 | hCoV-19/England/ALDP-A85634/2020\|EPI_ISL_610368\|20 | ···················· | ···················· | ····················· | ···················· | ···················· | ···················· |
| O | 2452 | hCoV-19/England/ALDP-A85E2D/2020\|EPI_ISL_610369\|20 | ···················· | ···················· | ····················· | ···················· | ···················· | ···················· |
| O | 2453 | hCoV-19/England/ALDP-A858A7/2020\|EPI_ISL_610370\|20 | ···················· | ···················· | ····················· | ···················· | ···················· | ···················· |
| O | 2454 | hCoV-19/England/ALDP-A85CBA/2020\|EPI_ISL_610372\|20 | ···················· | ···················· | ····················· | ···················· | ···················· | ···················· |
| O | 2455 | hCoV-19/England/ALDP-A859A4/2020\|EPI_ISL_610373\|20 | ···················· | ···················· | ····················· | ···················· | ···················· | ···················· |
| O | 2456 | hCoV-19/England/ALDP-A85ED2/2020\|EPI_ISL_610374\|20 | ···················· | ···················· | ····················· | ···················· | ···················· | ···················· |
| O | 2457 | hCoV-19/England/ALDP-A85B80/2020\|EPI_ISL_610375\|20 | ···················· | ···················· | ····················· | ···················· | ···················· | ···················· |
| O | 2458 | hCoV-19/England/ALDP-A85090/2020\|EPI_ISL_610376\|20 | ···················· | ···················· | ····················· | ···················· | ···················· | ···················· |
| O | 2459 | hCoV-19/England/ALDP-A84C60/2020\|EPI_ISL_610377\|20 | ···················· | ···················· | ····················· | ···················· | ···················· | ···················· |
| O | 2460 | hCoV-19/England/ALDP-A857B9/2020\|EPI_ISL_610378\|20 | ···················· | ···················· | ····················· | ···················· | ···················· | ···················· |
| O | 2461 | hCoV-19/England/ALDP-A8524F/2020\|EPI_ISL_610379\|20 | ···················· | ···················· | ····················· | ···················· | ···················· | ···················· |
| O | 2462 | hCoV-19/England/ALDP-A84978/2020\|EPI_ISL_610380\|20 | ···················· | ···················· | ····················· | ···················· | ···················· | ···················· |
| O | 2463 | hCoV-19/England/ALDP-A8535B/2020\|EPI_ISL_610381\|20 | ···················· | ···················· | ····················· | ···················· | ···················· | ···················· |
| O | 2464 | hCoV-19/England/ALDP-A8577D/2020\|EPI_ISL_610382\|20 | ···················· | ···················· | ····················· | ···················· | ···················· | ···················· |
| O | 2465 | hCoV-19/England/ALDP-A860EA/2020\|EPI_ISL_610383\|20 | ···················· | ···················· | ····················· | ···················· | ···················· | ···················· |
| O | 2466 | hCoV-19/England/ALDP-A85458/2020\|EPI_ISL_610384\|20 | ···················· | ···················· | ····················· | ···················· | ···················· | ···················· |
| O | 2467 | hCoV-19/England/ALDP-A85670/2020\|EPI_ISL_610385\|20 | ···················· | ···················· | ····················· | ···················· | ···················· | ···················· |
| O | 2468 | hCoV-19/England/ALDP-A84C24/2020\|EPI_ISL_610386\|20 | ···················· | ···················· | ····················· | ···················· | ···················· | ···················· |
| O | 2469 | hCoV-19/England/ALDP-A858B6/2020\|EPI_ISL_610387\|20 | ···················· | ···················· | ····················· | ···················· | ···················· | ···················· |
| O | 2470 | hCoV-19/England/ALDP-A84F76/2020\|EPI_ISL_610388\|20 | ···················· | ···················· | ····················· | ···················· | ···················· | ···················· |
| O | 2471 | hCoV-19/England/ALDP-A85BCC/2020\|EPI_ISL_610389\|20 | ···················· | ···················· | ····················· | ···················· | ···················· | ···················· |

| | | | | | | | | |
|---|---|---|---|---|---|---|---|---|
| O | 2472 | hCoV-19/England/ALDP-A84E3D/2020\|EPI_ISL_610390\|20 | ···················· | ···················· | ···················· | ···················· | ···················· | ···················· |
| O | 2473 | hCoV-19/England/ALDP-A84C7F/2020\|EPI_ISL_610391\|20 | ···················· | ···················· | ···················· | ···················· | ···················· | ···················· |
| O | 2474 | hCoV-19/England/ALDP-A8592C/2020\|EPI_ISL_610392\|20 | ···················· | ···················· | ···················· | ···················· | ···················· | ···················· |
| O | 2475 | hCoV-19/England/ALDP-A84FA3/2020\|EPI_ISL_610394\|20 | ···················· | ···················· | ···················· | ···················· | ···················· | ···················· |
| O | 2476 | hCoV-19/England/ALDP-A85C50/2020\|EPI_ISL_610395\|20 | ···················· | ···················· | ···················· | ···················· | ···················· | ···················· |
| O | 2477 | hCoV-19/England/ALDP-A85573/2020\|EPI_ISL_610397\|20 | ···················· | ···················· | ···················· | ···················· | ···················· | ···················· |
| O | 2478 | hCoV-19/England/ALDP-A85C7E/2020\|EPI_ISL_610399\|20 | ···················· | ···················· | ···················· | ···················· | ···················· | ···················· |
| O | 2479 | hCoV-19/Wales/PHWC-480541/2020\|EPI_ISL_612900\|2020 | ···················· | ···················· | ···················· | ···················· | ···················· | ···················· |
| O | 2480 | hCoV-19/Wales/PHWC-480550/2020\|EPI_ISL_612901\|2020 | ···················· | ···················· | ···················· | ···················· | ···················· | ···················· |
| O | 2481 | hCoV-19/Wales/PHWC-48056F/2020\|EPI_ISL_612902\|2020 | ···················· | ···················· | ···················· | ···················· | ···················· | ···················· |
| O | 2482 | hCoV-19/Wales/PHWC-48057E/2020\|EPI_ISL_612903\|2020 | ···················· | ···················· | ···················· | ···················· | ···················· | ···················· |
| O | 2483 | hCoV-19/Wales/PHWC-48058D/2020\|EPI_ISL_612904\|2020 | ···················· | ···················· | ···················· | ···················· | ···················· | ···················· |
| O | 2484 | hCoV-19/Wales/PHWC-4805BA/2020\|EPI_ISL_612905\|2020 | ···················· | ···················· | ···················· | ···················· | ···················· | ···················· |
| O | 2485 | hCoV-19/Wales/PHWC-4805C9/2020\|EPI_ISL_612906\|2020 | ···················· | ···················· | ···················· | ···················· | ···················· | ···················· |
| O | 2486 | hCoV-19/Wales/PHWC-4805D8/2020\|EPI_ISL_612907\|2020 | ···················· | ···················· | ···················· | ···················· | ···················· | ···················· |
| O | 2487 | hCoV-19/Wales/PHWC-4805E7/2020\|EPI_ISL_612908\|2020 | ···················· | ···················· | ···················· | ···················· | ···················· | ···················· |
| O | 2488 | hCoV-19/Wales/PHWC-4805F6/2020\|EPI_ISL_612909\|2020 | ···················· | ···················· | ···················· | ···················· | ···················· | ···················· |
| O | 2489 | hCoV-19/Wales/PHWC-480602/2020\|EPI_ISL_612910\|2020 | ···················· | ···················· | ···················· | ···················· | ···················· | ···················· |
| O | 2490 | hCoV-19/Wales/PHWC-480611/2020\|EPI_ISL_612911\|2020 | ···················· | ···················· | ···················· | ···················· | ···················· | ···················· |
| O | 2491 | hCoV-19/Wales/PHWC-480620/2020\|EPI_ISL_612912\|2020 | ···················· | ···················· | ···················· | ···················· | ···················· | ···················· |
| O | 2493 | hCoV-19/Wales/PHWC-48064E/2020\|EPI_ISL_612914\|2020 | ···················· | ···················· | ···················· | ···················· | ···················· | ···················· |
| O | 2494 | hCoV-19/Wales/PHWC-48066C/2020\|EPI_ISL_612915\|2020 | ···················· | ···················· | ···················· | ···················· | ···················· | ···················· |
| O | 2495 | hCoV-19/Wales/PHWC-48067B/2020\|EPI_ISL_612916\|2020 | ···················· | ···················· | ···················· | ···················· | ···················· | ···················· |
| O | 2496 | hCoV-19/Wales/PHWC-480699/2020\|EPI_ISL_612917\|2020 | ···················· | ···················· | ···················· | ···················· | ···················· | ···················· |
| O | 2497 | hCoV-19/Wales/PHWC-4806B7/2020\|EPI_ISL_612918\|2020 | ···················· | ···················· | ···················· | ···················· | ···················· | ···················· |
| O | 2498 | hCoV-19/Wales/PHWC-4806C6/2020\|EPI_ISL_612919\|2020 | ···················· | ···················· | ···················· | ···················· | ···················· | ···················· |
| O | 2499 | hCoV-19/Wales/PHWC-4806D5/2020\|EPI_ISL_612920\|2020 | ···················· | ···················· | ···················· | ···················· | ···················· | ···················· |
| O | 2500 | hCoV-19/Wales/PHWC-4806E4/2020\|EPI_ISL_612921\|2020 | ···················· | ···················· | ···················· | ···················· | ···················· | ···················· |
| O | 2501 | hCoV-19/Wales/PHWC-48070F/2020\|EPI_ISL_612922\|2020 | ···················· | ···················· | ···················· | ···················· | ··NN················ | ···················· |
| O | 2502 | hCoV-19/Wales/PHWC-48072D/2020\|EPI_ISL_612923\|2020 | ···················· | ···················· | ···················· | ···················· | ···················· | ···················· |
| O | 2503 | hCoV-19/Wales/PHWC-48073C/2020\|EPI_ISL_612924\|2020 | ···················· | ···················· | ···················· | ···················· | ·NN················· | ···················· |
| O | 2504 | hCoV-19/Wales/PHWC-48074B/2020\|EPI_ISL_612925\|2020 | ···················· | ···················· | ···················· | ···················· | ···················· | ···················· |
| O | 2505 | hCoV-19/Wales/PHWC-48075A/2020\|EPI_ISL_612926\|2020 | ···················· | ···················· | ···················· | ···················· | ···················· | ···················· |
| O | 2506 | hCoV-19/Wales/PHWC-480769/2020\|EPI_ISL_612927\|2020 | ···················· | ···················· | ···················· | ···················· | ···················· | ···················· |
| O | 2507 | hCoV-19/Wales/PHWC-480778/2020\|EPI_ISL_612928\|2020 | ···················· | ···················· | ···················· | ···················· | ···················· | ···················· |
| O | 2508 | hCoV-19/Wales/PHWC-480787/2020\|EPI_ISL_612929\|2020 | ···················· | ···················· | ···················· | ···················· | ···················· | ···················· |
| O | 2509 | hCoV-19/Wales/PHWC-480796/2020\|EPI_ISL_612930\|2020 | ···················· | ···················· | ···················· | ···················· | ···················· | ···················· |
| O | 2510 | hCoV-19/Wales/PHWC-4807A5/2020\|EPI_ISL_612931\|2020 | ···················· | ···················· | ···················· | ···················· | ···················· | ···················· |
| O | 2511 | hCoV-19/Wales/PHWC-4807C3/2020\|EPI_ISL_612932\|2020 | ···················· | ···················· | ···················· | ···················· | ···················· | ···················· |
| O | 2513 | hCoV-19/Wales/PHWC-4807F0/2020\|EPI_ISL_612934\|2020 | ···················· | ···················· | ···················· | ···················· | ···················· | ···················· |
| O | 2514 | hCoV-19/Wales/PHWC-48080C/2020\|EPI_ISL_612935\|2020 | ···················· | ···················· | ···················· | ···················· | ···················· | ···················· |
| O | 2516 | hCoV-19/Wales/PHWC-48082A/2020\|EPI_ISL_612937\|2020 | ···················· | ···················· | ···················· | ···················· | ···················· | ···················· |
| O | 2517 | hCoV-19/Wales/PHWC-480839/2020\|EPI_ISL_612938\|2020 | ···················· | ···················· | ···················· | ···················· | ···················· | ···················· |
| O | 2518 | hCoV-19/Wales/PHWC-480848/2020\|EPI_ISL_612939\|2020 | ···················· | ···················· | ···················· | ···················· | ···················· | ···················· |
| O | 2520 | hCoV-19/Wales/PHWC-480866/2020\|EPI_ISL_612941\|2020 | ···················· | ···················· | ···················· | ···················· | ···················· | ···················· |
| O | 2521 | hCoV-19/Wales/PHWC-480884/2020\|EPI_ISL_612942\|2020 | ···················· | ···················· | ···················· | ···················· | ···················· | ···················· |
| O | 2522 | hCoV-19/Wales/PHWC-4808A2/2020\|EPI_ISL_612943\|2020 | ···················· | ···················· | ···················· | ···················· | ···················· | ···················· |
| O | 2523 | hCoV-19/Wales/PHWC-4808B1/2020\|EPI_ISL_612944\|2020 | ···················· | ···················· | ···················· | ···················· | ···················· | ···················· |
| O | 2524 | hCoV-19/Wales/PHWC-4808C0/2020\|EPI_ISL_612945\|2020 | ···················· | ···················· | ···················· | ···················· | ···················· | ···················· |
| O | 2525 | hCoV-19/Wales/PHWC-4808DF/2020\|EPI_ISL_612946\|2020 | ···················· | ···················· | ···················· | ···················· | ···················· | ···················· |
| O | 2526 | hCoV-19/Wales/PHWC-4808EE/2020\|EPI_ISL_612947\|2020 | ···················· | ···················· | ···················· | ···················· | ···················· | ···················· |
| O | 2527 | hCoV-19/Wales/PHWC-4808FD/2020\|EPI_ISL_612948\|2020 | ···················· | ···················· | ···················· | ···················· | ···················· | ···················· |
| O | 2528 | hCoV-19/Wales/PHWC-480909/2020\|EPI_ISL_612949\|2020 | ···················· | ···················· | ···················· | ···················· | ···················· | ···················· |
| O | 2529 | hCoV-19/Wales/PHWC-480918/2020\|EPI_ISL_612950\|2020 | ···················· | ···················· | ···················· | ···················· | ···················· | ···················· |
| O | 2530 | hCoV-19/Wales/PHWC-480927/2020\|EPI_ISL_612951\|2020 | ···················· | ···················· | ···················· | ···················· | ···················· | ···················· |
| O | 2531 | hCoV-19/Wales/PHWC-480936/2020\|EPI_ISL_612952\|2020 | ···················· | ···················· | ···················· | ···················· | ···················· | ···················· |
| O | 2532 | hCoV-19/Wales/PHWC-480963/2020\|EPI_ISL_612953\|2020 | ···················· | ···················· | ···················· | ···················· | ···················· | ···················· |
| O | 2533 | hCoV-19/Wales/PHWC-480972/2020\|EPI_ISL_612954\|2020 | ···················· | ···················· | ···················· | ···················· | ···················· | ···················· |
| O | 2534 | hCoV-19/Wales/PHWC-480981/2020\|EPI_ISL_612955\|2020 | ···················· | ···················· | ···················· | ···················· | ···················· | ···················· |
| O | 2535 | hCoV-19/Wales/PHWC-480990/2020\|EPI_ISL_612956\|2020 | ···················· | ···················· | ···················· | ···················· | ···················· | ···················· |
| O | 2537 | hCoV-19/Wales/PHWC-4809DC/2020\|EPI_ISL_612958\|2020 | ···················· | ···················· | ···················· | ···················· | ···················· | ···················· |
| O | 2538 | hCoV-19/Wales/PHWC-4809EB/2020\|EPI_ISL_612959\|2020 | ···················· | ···················· | ···················· | ···················· | ···················· | ···················· |
| O | 2539 | hCoV-19/Wales/PHWC-480A15/2020\|EPI_ISL_612960\|2020 | ···················· | ···················· | ···················· | ···················· | ···················· | ···················· |
| O | 2540 | hCoV-19/Wales/PHWC-480A24/2020\|EPI_ISL_612961\|2020 | ···················· | ···················· | ···················· | ···················· | ···················· | ···················· |
| O | 2541 | hCoV-19/Wales/PHWC-480A42/2020\|EPI_ISL_612962\|2020 | ···················· | ···················· | ···················· | ···················· | ···················· | ···················· |
| O | 2542 | hCoV-19/Wales/PHWC-480A60/2020\|EPI_ISL_612963\|2020 | ···················· | ···················· | ···················· | ···················· | ···················· | ···················· |
| O | 2543 | hCoV-19/Wales/PHWC-480A7F/2020\|EPI_ISL_612964\|2020 | ···················· | ···················· | ···················· | ···················· | ···················· | ···················· |
| O | 2544 | hCoV-19/Wales/PHWC-480A8E/2020\|EPI_ISL_612965\|2020 | ···················· | ···················· | ···················· | ···················· | ···················· | ···················· |
| O | 2545 | hCoV-19/Wales/PHWC-480AAC/2020\|EPI_ISL_612966\|2020 | ···················· | ···················· | ···················· | ···················· | ···················· | ···················· |
| O | 2546 | hCoV-19/Wales/PHWC-480ABB/2020\|EPI_ISL_612967\|2020 | ···················· | ···················· | ···················· | ···················· | ···················· | ···················· |
| O | 2547 | hCoV-19/Wales/PHWC-480ACA/2020\|EPI_ISL_612968\|2020 | ···················· | ···················· | ···················· | ···················· | ···················· | ···················· |
| O | 2549 | hCoV-19/Wales/PHWC-480AE8/2020\|EPI_ISL_612970\|2020 | ···················· | ···················· | ···················· | ···················· | ···················· | ···················· |
| O | 2550 | hCoV-19/Wales/PHWC-480AF7/2020\|EPI_ISL_612971\|2020 | ···················· | ···················· | ···················· | ···················· | ···················· | ···················· |
| O | 2551 | hCoV-19/Wales/PHWC-480B03/2020\|EPI_ISL_612972\|2020 | ···················· | ···················· | ···················· | ···················· | ···················· | ···················· |
| O | 2552 | hCoV-19/Wales/PHWC-480B21/2020\|EPI_ISL_612973\|2020 | ···················· | ···················· | ···················· | ···················· | ···················· | ···················· |
| O | 2553 | hCoV-19/Wales/PHWC-480B30/2020\|EPI_ISL_612974\|2020 | ···················· | ···················· | ···················· | ···················· | ···················· | ···················· |
| O | 2556 | hCoV-19/Wales/PHWC-480B9A/2020\|EPI_ISL_612980\|2020 | ···················· | ···················· | ···················· | ···················· | ···················· | ···················· |
| O | 2557 | hCoV-19/Wales/PHWC-480BA9/2020\|EPI_ISL_612981\|2020 | ···················· | ···················· | ···················· | ···················· | ···················· | ···················· |
| O | 2558 | hCoV-19/Wales/PHWC-480BD6/2020\|EPI_ISL_612982\|2020 | ···················· | ···················· | ···················· | ···················· | ···················· | ···················· |
| O | 2559 | hCoV-19/Wales/PHWC-480BE5/2020\|EPI_ISL_612983\|2020 | ···················· | ···················· | ···················· | ···················· | ···················· | ···················· |

O 2560 hCoV-19/Wales/PHWC-480C4C/2020|EPI_ISL_612986|2020 ···················· ···················· ····················· ···················· ···················· ····················
O 2561 hCoV-19/India/DL-ILBS-279M/2020|EPI_ISL_605799|202 ···················· ···················· ····················· ···················· ···················· ····················
O 2562 hCoV-19/England/ALDP-A6FF44/2020|EPI_ISL_608000|20 ···················· ···················· ····················· ···················· ···················· ····················
O 2563 hCoV-19/England/ALDP-A6C5AF/2020|EPI_ISL_608001|20 ···················· ···················· ····················· ···················· ···················· ····················
O 2564 hCoV-19/England/ALDP-A6CB70/2020|EPI_ISL_608002|20 ···················· ···················· ····················· ···················· ···················· ····················
O 2565 hCoV-19/England/ALDP-A6D3C2/2020|EPI_ISL_608003|20 ···················· ···················· ····················· ···················· ···················· ····················
O 2566 hCoV-19/England/ALDP-A70872/2020|EPI_ISL_608004|20 ···················· ···················· ····················· ···················· ···················· ····················
O 2567 hCoV-19/England/ALDP-A701C2/2020|EPI_ISL_608005|20 ···················· ···················· ····················· ···················· ···················· ····················
O 2568 hCoV-19/England/ALDP-A6CF65/2020|EPI_ISL_608006|20 ···················· ···················· ····················· ···················· ···················· ····················
O 2569 hCoV-19/England/ALDP-A6D0CB/2020|EPI_ISL_608007|20 ···················· ···················· ····················· ···················· ···················· ····················
O 2570 hCoV-19/England/ALDP-A7010E/2020|EPI_ISL_608008|20 ···················· ·A·················· ····················· ···················· ···················· ····················
O 2571 hCoV-19/England/ALDP-A6D25C/2020|EPI_ISL_608009|20 ···················· ···················· ····················· ···················· ···················· ····················
O 2572 hCoV-19/England/ALDP-A6D3FF/2020|EPI_ISL_608010|20 ···················· ···················· ····················· ···················· ···················· ····················
O 2573 hCoV-19/England/ALDP-A6D34A/2020|EPI_ISL_608011|20 ···················· ···················· ····················· ···················· ···················· ····················
O 2574 hCoV-19/England/ALDP-A6CE86/2020|EPI_ISL_608012|20 ···················· ···················· ····················· ···················· ······T············· ····················
O 2575 hCoV-19/England/ALDP-A6D33B/2020|EPI_ISL_608013|20 ···················· ···················· ····················· ···················· ···················· ····················
O 2576 hCoV-19/England/ALDP-A6CE0E/2020|EPI_ISL_608014|20 ···················· ···················· ····················· ···················· ···················· ····················
O 2577 hCoV-19/England/ALDP-A6FF08/2020|EPI_ISL_608015|20 ···················· ···················· ····················· ···················· ···················· ····················
O 2578 hCoV-19/England/ALDP-A6CBCB/2020|EPI_ISL_608016|20 ···················· ···················· ····················· ···················· ···················· ····················
O 2579 hCoV-19/England/ALDP-A6D26B/2020|EPI_ISL_608017|20 ···················· ···················· ····················· ···················· ···················· ····················
O 2580 hCoV-19/England/ALDP-A6C81F/2020|EPI_ISL_608018|20 ···················· ···················· ····················· ···················· ···················· ····················
O 2581 hCoV-19/England/ALDP-A70942/2020|EPI_ISL_608019|20 ···················· ···················· ····················· ···················· ···················· ····················
O 2582 hCoV-19/England/ALDP-A70854/2020|EPI_ISL_608020|20 ···················· ·A·················· ····················· ···················· ···················· ····················
O 2583 hCoV-19/England/ALDP-A6CCAA/2020|EPI_ISL_608021|20 ···················· ···················· ····················· ···················· ···················· ····················
O 2584 hCoV-19/England/ALDP-A7002F/2020|EPI_ISL_608022|20 ···················· ···················· ····················· ···················· ···················· ····················
O 2585 hCoV-19/England/ALDP-A6C448/2020|EPI_ISL_608023|20 ···················· ···················· ····················· ···················· ······T············· ···············A····
O 2586 hCoV-19/England/ALDP-A6CD7A/2020|EPI_ISL_608024|20 ···················· ···················· ····················· ···················· ···················· ····················
O 2587 hCoV-19/England/ALDP-A6FEED/2020|EPI_ISL_608025|20 ···················· ···················· ····················· ···················· ···················· ····················
O 2588 hCoV-19/England/ALDP-A707B1/2020|EPI_ISL_608026|20 ···················· ···················· ····················· ···················· ···················· ····················
O 2589 hCoV-19/England/ALDP-A6C34B/2020|EPI_ISL_608028|20 ···················· ···················· ····················· ···················· ···················· ····················
O 2590 hCoV-19/England/ALDP-A6D2D4/2020|EPI_ISL_608029|20 ···················· ···················· ····················· ···················· ···················· ····················
O 2591 hCoV-19/England/ALDP-A6CD4D/2020|EPI_ISL_608030|20 ···················· ···················· ····················· ···················· ···················· ····················
O 2592 hCoV-19/England/ALDP-A6CB8F/2020|EPI_ISL_608033|20 ···················· ···················· ····················· ···················· ···················· ····················
O 2593 hCoV-19/England/ALDP-A701A4/2020|EPI_ISL_608034|20 ···················· ···················· ····················· ···················· ···················· ····················
O 2594 hCoV-19/England/ALDP-A6C387/2020|EPI_ISL_608035|20 ···················· ···················· ····················· ···················· ···················· ····················
O 2595 hCoV-19/England/ALDP-A6D799/2020|EPI_ISL_608036|20 ···················· ···················· ····················· ···················· ···················· ····················
O 2596 hCoV-19/England/ALDP-A6CF92/2020|EPI_ISL_608037|20 ···················· ···················· ····················· ···················· ···················· ····················
O 2597 hCoV-19/England/ALDP-A6D8D2/2020|EPI_ISL_608038|20 ···················· ···················· ····················· ···················· ···················· ····················
O 2598 hCoV-19/England/ALDP-A6D210/2020|EPI_ISL_608039|20 ···················· ···················· ····················· ···················· ···················· ····················
O 2599 hCoV-19/Scotland/CAMC-A570F1/2020|EPI_ISL_609200|2 ···················· ···················· ····················· ···················· ···················· ····················
O 2600 hCoV-19/Wales/CAMC-A5797E/2020|EPI_ISL_609201|2020 ···················· ···················· ····················· ···················· ···················· ····················
O 2601 hCoV-19/Scotland/QEUH-A5E616/2020|EPI_ISL_609202|2 ···················· ···················· ····················· ···················· ···················· ····················
O 2602 hCoV-19/Scotland/CAMC-A57352/2020|EPI_ISL_609203|2 ···················· ···················· ····················· ···················· ···················· ····················
O 2603 hCoV-19/Scotland/CAMC-A57B5A/2020|EPI_ISL_609204|2 ···················· ···················· ····················· ···················· ···················· ····················
O 2604 hCoV-19/Scotland/CAMC-A573DA/2020|EPI_ISL_609205|2 ···················· ···················· ····················· ···················· ···················· ····················
O 2605 hCoV-19/Wales/QEUH-A5DE6A/2020|EPI_ISL_609206|2020 ···················· ···················· ····················· ···················· ···················· ····················
O 2606 hCoV-19/Wales/ALDP-A5080F/2020|EPI_ISL_609207|2020 ···················· ···················· ····················· ···················· ···················· ····················
O 2607 hCoV-19/Wales/ALDP-A51130/2020|EPI_ISL_609208|2020 ···················· ···················· ····················· ···················· ···················· ····················
O 2608 hCoV-19/Wales/MILK-A46BD6/2020|EPI_ISL_609250|2020 ···················· ···················· ····················· ···················· ···················· ····················
O 2609 hCoV-19/Wales/MILK-A477A4/2020|EPI_ISL_609251|2020 ···················· ···················· ····················· ···················· ···················· ····················
O 2610 hCoV-19/Wales/MILK-A46E55/2020|EPI_ISL_609252|2020 ···················· ···················· ····················· ···················· ···················· ····················
O 2611 hCoV-19/Wales/MILK-A47B99/2020|EPI_ISL_609253|2020 ···················· ···················· ····················· ···················· ···················· ····················
O 2612 hCoV-19/Wales/MILK-A47EBE/2020|EPI_ISL_609254|2020 ···················· ···················· ····················· ···················· ···················· ····················
O 2613 hCoV-19/Wales/MILK-A47A9C/2020|EPI_ISL_609255|2020 ···················· ···················· ····················· ···················· ···················· ····················
O 2614 hCoV-19/Wales/MILK-A46B4F/2020|EPI_ISL_609256|2020 ···················· ···················· ····················· ···················· ···················· ····················
O 2615 hCoV-19/Wales/MILK-A46E64/2020|EPI_ISL_609257|2020 ···················· ···················· ····················· ···················· ···················· ····················
O 2616 hCoV-19/Wales/MILK-A47E09/2020|EPI_ISL_609259|2020 ···················· ···················· ····················· ···················· ···················· ····················
O 2617 hCoV-19/Wales/MILK-A46B7C/2020|EPI_ISL_609261|2020 ···················· ···················· ····················· ···················· ···················· ····················
O 2618 hCoV-19/Wales/MILK-A46BA9/2020|EPI_ISL_609262|2020 ···················· ···················· ····················· ···················· ···················· ····················
O 2619 hCoV-19/Wales/MILK-A470D6/2020|EPI_ISL_609263|2020 ···················· ···················· ····················· ···················· ···················· ····················
O 2620 hCoV-19/Wales/MILK-A47795/2020|EPI_ISL_609264|2020 ···················· ···················· ····················· ···················· ···················· ····················
O 2621 hCoV-19/Wales/MILK-A47B5D/2020|EPI_ISL_609265|2020 ···················· ···················· ····················· ···················· ···················· ····················
O 2622 hCoV-19/Wales/MILK-A47C1E/2020|EPI_ISL_609266|2020 ···················· ···················· ····················· ···················· ···················· ····················
O 2623 hCoV-19/Wales/MILK-A47E54/2020|EPI_ISL_609268|2020 ···················· ···················· ····················· ···················· ···················· ····················
O 2624 hCoV-19/Wales/MILK-A4748F/2020|EPI_ISL_609269|2020 ···················· ···················· ····················· ···················· ···················· ····················
O 2625 hCoV-19/Wales/MILK-A47BA8/2020|EPI_ISL_609274|2020 ···················· ···················· ····················· ···················· ···················· ····················
O 2626 hCoV-19/Wales/MILK-A477B3/2020|EPI_ISL_609275|2020 ···················· ···················· ····················· ···················· ···················· ····················
O 2627 hCoV-19/Wales/MILK-A463A1/2020|EPI_ISL_609290|2020 ···················· ···················· ····················· ···················· ···················· ····················
O 2628 hCoV-19/Wales/MILK-A46347/2020|EPI_ISL_609292|2020 ···················· ···················· ····················· ···················· ···················· ····················
O 2629 hCoV-19/Wales/MILK-A44D5A/2020|EPI_ISL_609293|2020 ···················· ···················· ····················· ···················· ···················· ····················
O 2630 hCoV-19/Wales/MILK-A46796/2020|EPI_ISL_609294|2020 ···················· ···················· ····················· ···················· ···················· ····················
O 2631 hCoV-19/Wales/MILK-A467F0/2020|EPI_ISL_609295|2020 ···················· ···················· ····················· ···················· ···················· ····················
O 2632 hCoV-19/Wales/MILK-A46417/2020|EPI_ISL_609296|2020 ···················· ···················· ····················· ···················· ···················· ····················
O 2633 hCoV-19/England/MILK-A45C3E/2020|EPI_ISL_609298|20 ···················· ···················· ····················· ···················· ···················· ····················
O 2634 hCoV-19/Wales/MILK-A44DA5/2020|EPI_ISL_609299|2020 ···················· ···················· ····················· ···················· ···················· ····················
O 2635 hCoV-19/England/CAMC-A41B62/2020|EPI_ISL_609331|20 ···················· ···················· ····················· ···················· ···················· ····················
O 2636 hCoV-19/England/CAMC-A41F48/2020|EPI_ISL_609332|20 ···················· ···················· ····················· ···················· ···················· ····················
O 2637 hCoV-19/England/CAMC-A418B6/2020|EPI_ISL_609333|20 ···················· ···················· ····················· ···················· ···················· ····················
O 2638 hCoV-19/England/CAMC-A4136A/2020|EPI_ISL_609334|20 ···················· ···················· ····················· ···················· ···················· ····················
O 2639 hCoV-19/England/CAMC-A41BDB/2020|EPI_ISL_609336|20 ···················· ···················· ····················· ···················· ···················· ····················

| | | | | | | | | |
|---|---|---|---|---|---|---|---|---|
| O | 2640 | hCoV-19/England/CAMC-A41EF0/2020\|EPI_ISL_609337\|20 | ···················· | ···················· | ···················· | ···················· | ···················· | ···················· |
| O | 2641 | hCoV-19/England/CAMC-A41713/2020\|EPI_ISL_609338\|20 | ···················· | ···················· | ···················· | ···················· | ···················· | ···················· |
| O | 2642 | hCoV-19/England/CAMC-A40E2E/2020\|EPI_ISL_609339\|20 | ···················· | ···················· | ···················· | ···················· | ···················· | ···················· |
| O | 2643 | hCoV-19/England/CAMC-A41555/2020\|EPI_ISL_609340\|20 | ···················· | ···················· | ···················· | ···················· | ···················· | ···················· |
| O | 2644 | hCoV-19/England/CAMC-A40D12/2020\|EPI_ISL_609341\|20 | ···················· | ···················· | ···················· | ···················· | ···················· | ···················· |
| O | 2645 | hCoV-19/England/CAMC-A417C8/2020\|EPI_ISL_609342\|20 | ···················· | ···················· | ···················· | ···················· | ···················· | ···················· |
| O | 2646 | hCoV-19/England/CAMC-A40E5B/2020\|EPI_ISL_609344\|20 | ···················· | ···················· | ···················· | ···················· | ···················· | ···················· |
| O | 2647 | hCoV-19/England/CAMC-A4125E/2020\|EPI_ISL_609345\|20 | ···················· | ···················· | ···················· | ···················· | ···················· | ···················· |
| O | 2648 | hCoV-19/England/CAMC-A41C50/2020\|EPI_ISL_609346\|20 | ···················· | ···················· | ···················· | ···················· | ···················· | ···················· |
| O | 2649 | hCoV-19/England/CAMC-A41E2D/2020\|EPI_ISL_609347\|20 | ···················· | ·A·················· | ···················· | ···················· | ···················· | ···················· |
| O | 2650 | hCoV-19/England/CAMC-A42071/2020\|EPI_ISL_609348\|20 | ···················· | ···················· | ···················· | ···················· | ···················· | ···················· |
| O | 2651 | hCoV-19/England/CAMC-A4140D/2020\|EPI_ISL_609349\|20 | ···················· | ···················· | ···················· | ···················· | ···················· | ···················· |
| O | 2652 | hCoV-19/England/CAMC-A4117F/2020\|EPI_ISL_609350\|20 | ···················· | ···················· | ···················· | ···················· | ···················· | ···················· |
| O | 2653 | hCoV-19/England/CAMC-A41A0B/2020\|EPI_ISL_609351\|20 | ···················· | ···················· | ···················· | ···················· | ···················· | ···················· |
| O | 2654 | hCoV-19/England/CAMC-A4124F/2020\|EPI_ISL_609352\|20 | ···················· | ···················· | ···················· | ···················· | ···················· | ···················· |
| O | 2655 | hCoV-19/England/CAMC-A41E5A/2020\|EPI_ISL_609353\|20 | ···················· | ···················· | ···················· | ···················· | ···················· | ···················· |
| O | 2656 | hCoV-19/England/CAMC-A4175F/2020\|EPI_ISL_609354\|20 | ···················· | ·A·················· | ···················· | ···················· | ···················· | ··A················· |
| O | 2657 | hCoV-19/England/CAMC-A41A47/2020\|EPI_ISL_609356\|20 | ···················· | ···················· | ···················· | ···················· | ···················· | ···················· |
| O | 2658 | hCoV-19/England/CAMC-A41670/2020\|EPI_ISL_609357\|20 | ···················· | ···················· | ···················· | ···················· | ···················· | ···················· |
| O | 2659 | hCoV-19/England/CAMC-A414D0/2020\|EPI_ISL_609358\|20 | ···················· | ···················· | ···················· | ···················· | ···················· | ···················· |
| O | 2660 | hCoV-19/England/CAMC-A40E6A/2020\|EPI_ISL_609359\|20 | ···················· | ···················· | ···················· | ···················· | ···················· | ···················· |
| O | 2661 | hCoV-19/England/CAMC-A422A8/2020\|EPI_ISL_609360\|20 | ···················· | ···················· | ···················· | ···················· | ···················· | ···················· |
| O | 2662 | hCoV-19/England/CAMC-A40E79/2020\|EPI_ISL_609361\|20 | ···················· | ···················· | ···················· | ···················· | ···················· | ···················· |
| O | 2663 | hCoV-19/England/CAMC-A412B8/2020\|EPI_ISL_609362\|20 | ···················· | ·A·················· | ···················· | ···················· | ···················· | ···················· |
| O | 2664 | hCoV-19/England/CAMC-A4235A/2020\|EPI_ISL_609363\|20 | ···················· | ···················· | ···················· | ···················· | ···················· | ···················· |
| O | 2665 | hCoV-19/England/CAMC-A418A7/2020\|EPI_ISL_609364\|20 | ···················· | ···················· | ···················· | ···················· | ···················· | ···················· |
| O | 2666 | hCoV-19/England/CAMC-A41E4B/2020\|EPI_ISL_609365\|20 | ···················· | ···················· | ···················· | ···················· | ···················· | ···················· |
| O | 2667 | hCoV-19/England/CAMC-A415EC/2020\|EPI_ISL_609366\|20 | ···················· | ···················· | ···················· | ···················· | ···················· | ···················· |
| O | 2668 | hCoV-19/England/CAMC-A414A3/2020\|EPI_ISL_609367\|20 | ···················· | ···················· | ···················· | ···················· | ···················· | ···················· |
| O | 2669 | hCoV-19/England/CAMC-A41E78/2020\|EPI_ISL_609368\|20 | ···················· | ·A·················· | ···················· | ···················· | ···················· | ··A················· |
| O | 2670 | hCoV-19/England/CAMC-A410CD/2020\|EPI_ISL_609369\|20 | ···················· | ···················· | ···················· | ···················· | ···················· | ···················· |
| O | 2671 | hCoV-19/Russia/CRIE300223/2020\|EPI_ISL_610238\|2020 | ···················· | ···················· | ···················· | ···················· | ···················· | ···················· |
| O | 2672 | hCoV-19/Russia/CRIE7158978/2020\|EPI_ISL_610239\|202 | ···················· | ···················· | ···················· | ···················· | ···················· | ···················· |
| O | 2673 | hCoV-19/Russia/CRIE7159118/2020\|EPI_ISL_610240\|202 | ···················· | ···················· | ···················· | ···················· | ···················· | ···················· |
| O | 2674 | hCoV-19/Russia/CRIE7171114/2020\|EPI_ISL_610241\|202 | ···················· | ···················· | ···················· | ···················· | ···················· | ···················· |
| O | 2675 | hCoV-19/Russia/CRIE7175362/2020\|EPI_ISL_610242\|202 | ···················· | ···················· | ···················· | ···················· | ···················· | ···················· |
| O | 2676 | hCoV-19/Russia/CRIE7158368/2020\|EPI_ISL_610243\|202 | ···················· | ···················· | ·········T·········· | ···················· | ···················· | ···················· |
| O | 2677 | hCoV-19/Russia/CRIE7249896/2020\|EPI_ISL_610244\|202 | ···················· | ···················· | ···················· | ···················· | ···················· | ···················· |
| O | 2678 | hCoV-19/Germany/BY-ER-1/2020\|EPI_ISL_610249\|2020-0 | ···················· | ···················· | ···················· | ···················· | ···················· | ···················· |
| O | 2679 | hCoV-19/USA/NM-UNM-00792/2020\|EPI_ISL_610258\|2020- | ···················· | ···················· | ···················· | ···················· | ···················· | ···················· |
| O | 2680 | hCoV-19/USA/NM-UNM-00793/2020\|EPI_ISL_610259\|2020- | ···················· | ···················· | ···················· | ···················· | ···················· | ···················· |
| O | 2681 | hCoV-19/USA/NM-UNM-00811/2020\|EPI_ISL_610263\|2020- | ···················· | ···················· | ···················· | ···················· | ···················· | ···················· |
| O | 2682 | hCoV-19/USA/NM-UNM-00812/2020\|EPI_ISL_610264\|2020- | ···················· | ···················· | ···················· | ···················· | ···················· | ···················· |
| O | 2683 | hCoV-19/USA/NM-UNM-00854/2020\|EPI_ISL_610280\|2020- | ···················· | ···················· | ···················· | ···················· | ···················· | ···················· |
| O | 2684 | hCoV-19/USA/NM-UNM-00855/2020\|EPI_ISL_610281\|2020- | ···················· | ···················· | ···················· | ···················· | ···················· | ···················· |
| O | 2685 | hCoV-19/USA/NM-UNM-00860/2020\|EPI_ISL_610282\|2020- | ···················· | ···················· | ···················· | ···················· | ···················· | ···················· |
| O | 2686 | hCoV-19/USA/WA-UW-16037/2020\|EPI_ISL_610283\|2020-0 | ···················· | ···················· | ···················· | ···················· | ···················· | ···················· |
| O | 2687 | hCoV-19/USA/WA-UW-16407/2020\|EPI_ISL_610284\|2020-0 | ···················· | ···················· | ···················· | ···················· | ···················· | ···················· |
| O | 2688 | hCoV-19/USA/WA-UW-13894/2020\|EPI_ISL_610285\|2020-0 | ···················· | ···················· | ···················· | ···················· | ···················· | ···················· |
| O | 2689 | hCoV-19/England/ALDP-A85F66/2020\|EPI_ISL_610286\|20 | ···················· | ···················· | ···················· | ···················· | ···················· | ···················· |
| O | 2690 | hCoV-19/England/ALDP-A857D7/2020\|EPI_ISL_610287\|20 | ···················· | ···················· | ···················· | ···················· | ···················· | ···················· |
| O | 2691 | hCoV-19/England/ALDP-A85E4B/2020\|EPI_ISL_610288\|20 | ···················· | ···················· | ···················· | ···················· | ···················· | ···················· |
| O | 2692 | hCoV-19/England/ALDP-A84AFD/2020\|EPI_ISL_610289\|20 | ···················· | ···················· | ···················· | ···················· | ···················· | ···················· |
| O | 2693 | hCoV-19/England/ALDP-A85EF0/2020\|EPI_ISL_610290\|20 | ···················· | ···················· | ···················· | ···················· | ···················· | ···················· |
| O | 2694 | hCoV-19/England/ALDP-A84B54/2020\|EPI_ISL_610291\|20 | ···················· | ···················· | ···················· | ···················· | ···················· | ···················· |
| O | 2695 | hCoV-19/England/ALDP-A853C4/2020\|EPI_ISL_610292\|20 | ···················· | ···················· | ···················· | ···················· | ···················· | ···················· |
| O | 2696 | hCoV-19/England/ALDP-A85E5A/2020\|EPI_ISL_610293\|20 | ···················· | ···················· | ···················· | ···················· | ···················· | ···················· |
| O | 2697 | hCoV-19/England/ALDP-A85722/2020\|EPI_ISL_610294\|20 | ···················· | ···················· | ···················· | ···················· | ···················· | ···················· |
| O | 2698 | hCoV-19/England/ALDP-A8550A/2020\|EPI_ISL_610295\|20 | ···················· | ···················· | ···················· | ···················· | ···················· | ···················· |
| O | 2699 | hCoV-19/England/ALDP-A8582F/2020\|EPI_ISL_610297\|20 | ···················· | ···················· | ···················· | ···················· | ···················· | ···················· |
| O | 2700 | hCoV-19/England/ALDP-A84FFE/2020\|EPI_ISL_610299\|20 | ···················· | ···················· | ···················· | ···················· | ···················· | ···················· |
| O | 2701 | hCoV-19/England/ALDP-A853E2/2020\|EPI_ISL_610315\|20 | ···················· | ···················· | ···················· | ···················· | ···················· | ···················· |
| O | 2702 | hCoV-19/England/ALDP-A85EE1/2020\|EPI_ISL_610316\|20 | ···················· | ···················· | ···················· | ···················· | ···················· | ···················· |
| O | 2703 | hCoV-19/England/ALDP-A850DC/2020\|EPI_ISL_610318\|20 | ···················· | ···················· | ···················· | ···················· | ···················· | ···················· |
| O | 2704 | hCoV-19/England/ALDP-A86026/2020\|EPI_ISL_610319\|20 | ···················· | ···················· | ···················· | ···················· | ···················· | ···················· |
| O | 2705 | hCoV-19/England/ALDP-A853F1/2020\|EPI_ISL_610322\|20 | ···················· | ···················· | ···················· | ···················· | ···················· | ···················· |
| O | 2706 | hCoV-19/England/ALDP-A86062/2020\|EPI_ISL_610323\|20 | ···················· | ···················· | ···················· | ···················· | ···················· | ···················· |
| O | 2707 | hCoV-19/England/ALDP-A85528/2020\|EPI_ISL_610324\|20 | ···················· | ···················· | ···················· | ···················· | ···················· | ···················· |
| O | 2708 | hCoV-19/England/ALDP-A84E88/2020\|EPI_ISL_610325\|20 | ···················· | ···················· | ···················· | ···················· | ···················· | ···················· |
| O | 2709 | hCoV-19/England/ALDP-A849C3/2020\|EPI_ISL_610326\|20 | ···················· | ···················· | ···················· | ···················· | ···················· | ···················· |
| O | 2710 | hCoV-19/England/ALDP-A85388/2020\|EPI_ISL_610327\|20 | ···················· | ···················· | ···················· | ···················· | ···················· | ···················· |
| O | 2711 | hCoV-19/England/ALDP-A84DF4/2020\|EPI_ISL_610328\|20 | ···················· | ···················· | ···················· | ···················· | ···················· | ···················· |
| O | 2712 | hCoV-19/England/ALDP-A85CE7/2020\|EPI_ISL_610329\|20 | ···················· | ···················· | ···················· | ···················· | ···················· | ···················· |
| O | 2713 | hCoV-19/England/ALDP-A8492D/2020\|EPI_ISL_610330\|20 | ···················· | ···················· | ···················· | ···················· | ···················· | ···················· |
| O | 2714 | hCoV-19/England/ALDP-A85C05/2020\|EPI_ISL_610331\|20 | ···················· | ···················· | ···················· | ···················· | ···················· | ···················· |
| O | 2715 | hCoV-19/England/ALDP-A84EF1/2020\|EPI_ISL_610333\|20 | ···················· | ···················· | ···················· | ···················· | ···················· | ···················· |
| O | 2716 | hCoV-19/England/ALDP-A84B45/2020\|EPI_ISL_610335\|20 | ···················· | ···················· | ···················· | ···················· | ···················· | ···················· |
| O | 2717 | hCoV-19/England/ALDP-A85A92/2020\|EPI_ISL_610336\|20 | ···················· | ···················· | ···················· | ···················· | ···················· | ···················· |
| O | 2718 | hCoV-19/England/ALDP-A85977/2020\|EPI_ISL_610337\|20 | ···················· | ···················· | ···················· | ···················· | ···················· | ···················· |
| O | 2719 | hCoV-19/England/ALDP-A84CE8/2020\|EPI_ISL_610338\|20 | ···················· | ···················· | ···················· | ···················· | ···················· | ···················· |

| | | | | | | | | | |
|---|---|---|---|---|---|---|---|---|---|
| O | 2720 | hCoV-19/England/ALDP-A85A83/2020\|EPI_ISL_610339\|20 | ···················· | ···················· | ···················· | ···················· | ···················· | ···················· | |
| O | 2721 | hCoV-19/England/ALDP-A85018/2020\|EPI_ISL_610340\|20 | ···················· | ···················· | ···················· | ···················· | ···················· | ···················· | |
| O | 2722 | hCoV-19/England/ALDP-A8579B/2020\|EPI_ISL_610341\|20 | ···················· | ·A·················· | ···················· | ···················· | ···················· | ···················· | |
| O | 2723 | hCoV-19/England/ALDP-A8518E/2020\|EPI_ISL_610342\|20 | ···················· | ·A·················· | ···················· | ···················· | ···················· | ···················· | |
| O | 2724 | hCoV-19/England/ALDP-A8586B/2020\|EPI_ISL_610343\|20 | ···················· | ···················· | ···················· | ···················· | ···················· | ···················· | |
| O | 2725 | hCoV-19/England/ALDP-A85449/2020\|EPI_ISL_610346\|20 | ···················· | ···················· | ···················· | ···················· | ···················· | ···················· | |
| O | 2726 | hCoV-19/England/ALDP-A852A9/2020\|EPI_ISL_610347\|20 | ···················· | ···················· | ···················· | ···················· | ···················· | ···················· | |
| O | 2727 | hCoV-19/England/ALDP-A854A3/2020\|EPI_ISL_610348\|20 | ···················· | ···················· | ···················· | ···················· | ···················· | ···················· | |
| O | 2728 | hCoV-19/England/ALDP-A85B35/2020\|EPI_ISL_610349\|20 | ···················· | ···················· | ···················· | ···················· | ···················· | ···················· | |
| O | 2729 | hCoV-19/England/ALDP-A8583E/2020\|EPI_ISL_610350\|20 | ···················· | ···················· | ···················· | ···················· | ···················· | ···················· | |
| O | 2730 | hCoV-19/England/ALDP-A84B18/2020\|EPI_ISL_610351\|20 | ···················· | ···················· | ···················· | ···················· | ···················· | ···················· | |
| O | 2731 | hCoV-19/England/ALDP-A84C42/2020\|EPI_ISL_610352\|20 | ···················· | ···················· | ···················· | ···················· | ···················· | ···················· | |
| O | 2732 | hCoV-19/England/ALDP-A860DB/2020\|EPI_ISL_610353\|20 | ···················· | ···················· | ···················· | ···················· | ···················· | ···················· | |
| O | 2733 | hCoV-19/England/ALDP-A857AA/2020\|EPI_ISL_610354\|20 | ···················· | ···················· | ···················· | ···················· | ···················· | ···················· | |
| O | 2734 | hCoV-19/England/ALDP-A859E0/2020\|EPI_ISL_610355\|20 | ···················· | ···················· | ···················· | ···················· | ···················· | ···················· | |
| O | 2735 | hCoV-19/England/ALDP-A84BEB/2020\|EPI_ISL_610357\|20 | ···················· | ···················· | ···················· | ···················· | ···················· | ···················· | |
| O | 2736 | hCoV-19/England/ALDP-A84ADF/2020\|EPI_ISL_610358\|20 | ···················· | ···················· | ···················· | ···················· | ···················· | ···················· | |
| O | 2737 | hCoV-19/USA/UT-UPHL-201026960/2020\|EPI_ISL_611501\| | ···················· | ·A·················· | ···················· | ···················· | ···················· | ···················· | |
| O | 2738 | hCoV-19/USA/UT-UPHL-201012055/2020\|EPI_ISL_611502\| | ···················· | ·A·················· | ···················· | ···················· | ···················· | ···················· | |
| O | 2739 | hCoV-19/USA/UT-UPHL-201004279/2020\|EPI_ISL_611503\| | ···················· | ·A·················· | ···················· | ···················· | ···················· | ···················· | |
| O | 2740 | hCoV-19/USA/UT-UPHL-201003755/2020\|EPI_ISL_611504\| | ···················· | ·A·················· | ···················· | ···················· | ···················· | ···················· | |
| O | 2741 | hCoV-19/USA/UT-UPHL-201027253/2020\|EPI_ISL_611506\| | ···················· | ·A·················· | ···················· | ···················· | ···················· | ···················· | |
| O | 2742 | hCoV-19/USA/UT-UPHL-201031539/2020\|EPI_ISL_611513\| | ···················· | ···················· | ···················· | ···················· | ···················· | ···················· | |
| O | 2743 | hCoV-19/Wales/PHWC-47FD01/2020\|EPI_ISL_612800\|2020 | ···················· | ···················· | ···················· | ···················· | ···················· | ···················· | |
| O | 2744 | hCoV-19/Wales/PHWC-47FD10/2020\|EPI_ISL_612801\|2020 | ···················· | ···················· | ···················· | ···················· | ···················· | ···················· | |
| O | 2745 | hCoV-19/Wales/PHWC-47FD2F/2020\|EPI_ISL_612802\|2020 | ···················· | ···················· | ···················· | ···················· | ···················· | ···················· | |
| O | 2746 | hCoV-19/Wales/PHWC-47FD3E/2020\|EPI_ISL_612803\|2020 | ···················· | ···················· | ···················· | ···················· | ···················· | ···················· | |
| O | 2747 | hCoV-19/Wales/PHWC-47FD4D/2020\|EPI_ISL_612804\|2020 | ···················· | ···················· | ···················· | ···················· | ···················· | ···················· | |
| O | 2748 | hCoV-19/Wales/PHWC-47FD6B/2020\|EPI_ISL_612805\|2020 | ···················· | ···················· | ···················· | ···················· | ···················· | ···················· | |
| O | 2749 | hCoV-19/Wales/PHWC-47FD7A/2020\|EPI_ISL_612806\|2020 | ···················· | ···················· | ···················· | ···················· | ···················· | ···················· | |
| O | 2750 | hCoV-19/Wales/PHWC-47FD89/2020\|EPI_ISL_612807\|2020 | ···················· | ···················· | ···················· | ···················· | ···················· | ···················· | |
| O | 2751 | hCoV-19/Wales/PHWC-47FD98/2020\|EPI_ISL_612808\|2020 | ···················· | ···················· | ···················· | ···················· | ···················· | ···················· | |
| O | 2752 | hCoV-19/Wales/PHWC-47FDA7/2020\|EPI_ISL_612809\|2020 | ···················· | ···················· | ···················· | ···················· | ···················· | ···················· | |
| O | 2753 | hCoV-19/Wales/PHWC-47FDC5/2020\|EPI_ISL_612810\|2020 | ···················· | ···················· | ···················· | ···················· | ···················· | ···················· | |
| O | 2754 | hCoV-19/Wales/PHWC-47FDD4/2020\|EPI_ISL_612811\|2020 | ···················· | ···················· | ···················· | ···················· | ···················· | ···················· | |
| O | 2755 | hCoV-19/Wales/PHWC-47FDE3/2020\|EPI_ISL_612812\|2020 | ···················· | ···················· | ···················· | ···················· | ···················· | ···················· | |
| O | 2756 | hCoV-19/Wales/PHWC-47FDF2/2020\|EPI_ISL_612813\|2020 | ···················· | ···················· | ···················· | ···················· | ···················· | ···················· | |
| O | 2757 | hCoV-19/Wales/PHWC-47FE0E/2020\|EPI_ISL_612814\|2020 | ···················· | ···················· | ···················· | ···················· | ···················· | ···················· | |
| O | 2758 | hCoV-19/Wales/PHWC-47FE1D/2020\|EPI_ISL_612815\|2020 | ···················· | ···················· | ···················· | ···················· | ···················· | ···················· | |
| O | 2759 | hCoV-19/Wales/PHWC-47FE2C/2020\|EPI_ISL_612816\|2020 | ···················· | ···················· | ···················· | ···················· | NNN················· | ···················· | 1 |
| O | 2760 | hCoV-19/Wales/PHWC-47FE3B/2020\|EPI_ISL_612817\|2020 | ···················· | ···················· | ···················· | ···················· | ···················· | ···················· | |
| O | 2761 | hCoV-19/Wales/PHWC-47FE4A/2020\|EPI_ISL_612818\|2020 | ···················· | ···················· | ···················· | ···················· | ···················· | ···················· | |
| O | 2762 | hCoV-19/Wales/PHWC-47FE59/2020\|EPI_ISL_612819\|2020 | ···················· | ···················· | ···················· | ···················· | ···················· | ···················· | |
| O | 2763 | hCoV-19/Wales/PHWC-47FE68/2020\|EPI_ISL_612820\|2020 | ···················· | ···················· | ···················· | ···················· | ···················· | ···················· | |
| O | 2764 | hCoV-19/Wales/PHWC-47FE77/2020\|EPI_ISL_612821\|2020 | ···················· | ···················· | ···················· | ···················· | ···················· | ···················· | |
| O | 2765 | hCoV-19/Wales/PHWC-47FE86/2020\|EPI_ISL_612822\|2020 | ···················· | ···················· | ···················· | ···················· | ···················· | ···················· | |
| O | 2766 | hCoV-19/Wales/PHWC-47FEA4/2020\|EPI_ISL_612823\|2020 | ···················· | ···················· | ···················· | ···················· | ···················· | ···················· | |
| O | 2767 | hCoV-19/Wales/PHWC-47FEB3/2020\|EPI_ISL_612824\|2020 | ···················· | ···················· | ···················· | ···················· | ···················· | ···················· | |
| O | 2769 | hCoV-19/Wales/PHWC-47FED1/2020\|EPI_ISL_612826\|2020 | ···················· | ···················· | ···················· | ···················· | ···················· | ···················· | |
| O | 2770 | hCoV-19/Wales/PHWC-47FEE0/2020\|EPI_ISL_612827\|2020 | ···················· | ···················· | ···················· | ···················· | ···················· | ···················· | |
| O | 2771 | hCoV-19/Wales/PHWC-47FEFF/2020\|EPI_ISL_612828\|2020 | ···················· | ···················· | ···················· | ···················· | ···················· | ···················· | |
| O | 2772 | hCoV-19/Wales/PHWC-47FF0B/2020\|EPI_ISL_612829\|2020 | ···················· | ···················· | ···················· | ···················· | ···················· | ···················· | |
| O | 2773 | hCoV-19/Wales/PHWC-47FF1A/2020\|EPI_ISL_612830\|2020 | ···················· | ···················· | ···················· | ···················· | ···················· | ···················· | |
| O | 2774 | hCoV-19/Wales/PHWC-47FF29/2020\|EPI_ISL_612831\|2020 | ···················· | ···················· | ···················· | ···················· | ···················· | ···················· | |
| O | 2775 | hCoV-19/Wales/PHWC-47FF38/2020\|EPI_ISL_612832\|2020 | ···················· | ···················· | ···················· | ···················· | ···················· | ···················· | |
| O | 2776 | hCoV-19/Wales/PHWC-47FF65/2020\|EPI_ISL_612833\|2020 | ···················· | ···················· | ···················· | ···················· | ···················· | ···················· | |
| O | 2777 | hCoV-19/Wales/PHWC-47FF74/2020\|EPI_ISL_612834\|2020 | ···················· | ···················· | ···················· | ···················· | ···················· | ···················· | |
| O | 2778 | hCoV-19/Wales/PHWC-47FF83/2020\|EPI_ISL_612835\|2020 | ···················· | ···················· | ···················· | ···················· | ···················· | ···················· | |
| O | 2779 | hCoV-19/Wales/PHWC-47FF92/2020\|EPI_ISL_612836\|2020 | ···················· | ···················· | ···················· | ···················· | ···················· | ···················· | |
| O | 2780 | hCoV-19/Wales/PHWC-47FFA1/2020\|EPI_ISL_612837\|2020 | ···················· | ···················· | ···················· | ···················· | ···················· | ···················· | |
| O | 2781 | hCoV-19/Wales/PHWC-47FFCF/2020\|EPI_ISL_612838\|2020 | ···················· | ···················· | ···················· | ···················· | ···················· | ···················· | |
| O | 2782 | hCoV-19/Wales/PHWC-47FFDE/2020\|EPI_ISL_612839\|2020 | ···················· | ···················· | ···················· | ···················· | ···················· | ···················· | |
| O | 2783 | hCoV-19/Wales/PHWC-47FFED/2020\|EPI_ISL_612840\|2020 | ···················· | ···················· | ···················· | ···················· | NNN················· | ···················· | 1 |
| O | 2784 | hCoV-19/Wales/PHWC-47FFFC/2020\|EPI_ISL_612841\|2020 | ···················· | ···················· | ···················· | ···················· | ···················· | ···················· | |
| O | 2785 | hCoV-19/Wales/PHWC-480004/2020\|EPI_ISL_612842\|2020 | ···················· | ···················· | ···················· | ···················· | ···················· | ···················· | |
| O | 2786 | hCoV-19/Wales/PHWC-480031/2020\|EPI_ISL_612843\|2020 | ···················· | ···················· | ···················· | ···················· | ···················· | ···················· | |
| O | 2787 | hCoV-19/Wales/PHWC-480040/2020\|EPI_ISL_612844\|2020 | ···················· | ···················· | ···················· | ···················· | ···················· | ···················· | |
| O | 2789 | hCoV-19/Wales/PHWC-48007D/2020\|EPI_ISL_612846\|2020 | ···················· | ···················· | ···················· | ···················· | ···················· | ···················· | |
| O | 2790 | hCoV-19/Wales/PHWC-48009B/2020\|EPI_ISL_612847\|2020 | ···················· | ···················· | ···················· | ···················· | ···················· | ···················· | |
| O | 2793 | hCoV-19/Wales/PHWC-4800D7/2020\|EPI_ISL_612850\|2020 | ···················· | ···················· | ···················· | ···················· | ···················· | ···················· | |
| O | 2794 | hCoV-19/Wales/PHWC-4800E6/2020\|EPI_ISL_612851\|2020 | ···················· | ···················· | ···················· | ···················· | ···················· | ···················· | |
| O | 2795 | hCoV-19/Wales/PHWC-4800F5/2020\|EPI_ISL_612852\|2020 | ···················· | ···················· | ···················· | ···················· | ···················· | ···················· | |
| O | 2796 | hCoV-19/Wales/PHWC-48012F/2020\|EPI_ISL_612853\|2020 | ···················· | ···················· | ···················· | ···················· | ···················· | ···················· | |
| O | 2797 | hCoV-19/Wales/PHWC-48013E/2020\|EPI_ISL_612854\|2020 | ···················· | ···················· | ···················· | ···················· | ···················· | ···················· | |
| O | 2798 | hCoV-19/Wales/PHWC-48014D/2020\|EPI_ISL_612855\|2020 | ···················· | ···················· | ···················· | ···················· | ···················· | ···················· | |
| O | 2799 | hCoV-19/Wales/PHWC-480B6D/2020\|EPI_ISL_612977\|2020 | ···················· | ···················· | ···················· | ···················· | ···················· | ···················· | |
| O | 2800 | hCoV-19/Wales/PHWC-480B7C/2020\|EPI_ISL_612978\|2020 | ···················· | ···················· | ···················· | ···················· | ···················· | ···················· | |
| O | 2801 | hCoV-19/Wales/PHWC-480B8B/2020\|EPI_ISL_612979\|2020 | ···················· | ···················· | ···················· | ···················· | ···················· | ···················· | |
| O | 2802 | hCoV-19/Wales/PHWC-480C00/2020\|EPI_ISL_612984\|2020 | ···················· | ···················· | ···················· | ···················· | ···················· | ···················· | |
| O | 2803 | hCoV-19/Wales/PHWC-480C2E/2020\|EPI_ISL_612985\|2020 | ···················· | ···················· | ···················· | ···················· | ···················· | ···················· | |

| | | | | | | | |
|---|---|---|---|---|---|---|---|
| O | 2804 hCoV-19/Wales/PHWC-480C5B/2020\|EPI_ISL_612987\|2020 | ···················· | ···················· | ···················· | ···················· | ···················· | ···················· |
| O | 2805 hCoV-19/Wales/PHWC-480C6A/2020\|EPI_ISL_612988\|2020 | ···················· | ···················· | ···················· | ···················· | ···················· | ···················· |
| O | 2806 hCoV-19/Wales/PHWC-480C88/2020\|EPI_ISL_612989\|2020 | ···················· | ···················· | ···················· | ···················· | ···················· | ···················· |
| O | 2807 hCoV-19/Wales/PHWC-480CA6/2020\|EPI_ISL_612990\|2020 | ···················· | ···················· | ···················· | ···················· | ···················· | ···················· |
| O | 2808 hCoV-19/Wales/PHWC-480CC4/2020\|EPI_ISL_612991\|2020 | ···················· | ···················· | ···················· | ···················· | ···················· | ···················· |
| O | 2809 hCoV-19/Wales/PHWC-480CD3/2020\|EPI_ISL_612992\|2020 | ···················· | ···················· | ···················· | ···················· | ···················· | ···················· |
| O | 2810 hCoV-19/Wales/PHWC-480CE2/2020\|EPI_ISL_612993\|2020 | ···················· | ···················· | ···················· | ···················· | ···················· | ···················· |
| O | 2811 hCoV-19/Wales/PHWC-480D0D/2020\|EPI_ISL_612994\|2020 | ···················· | ···················· | ···················· | ···················· | ···················· | ···················· |
| O | 2812 hCoV-19/Wales/PHWC-480D1C/2020\|EPI_ISL_612995\|2020 | ···················· | ···················· | ···················· | ···················· | ···················· | ···················· |
| O | 2813 hCoV-19/Wales/PHWC-480D2B/2020\|EPI_ISL_612996\|2020 | ···················· | ···················· | ···················· | ···················· | ···················· | ···················· |
| O | 2814 hCoV-19/Wales/PHWC-480D3A/2020\|EPI_ISL_612997\|2020 | ···················· | ···················· | ···················· | ···················· | ···················· | ···················· |
| O | 2815 hCoV-19/Wales/PHWC-480D58/2020\|EPI_ISL_612998\|2020 | ···················· | ···················· | ···················· | ···················· | ···················· | ···················· |
| O | 2816 hCoV-19/Wales/PHWC-480D94/2020\|EPI_ISL_612999\|2020 | ···················· | ···················· | ···················· | ···················· | ···················· | ···················· |
| O | 2817 hCoV-19/England/MILK-A7C1A8/2020\|EPI_ISL_607400\|20 | ···················· | ···················· | ···················· | ···················· | ···················· | ···················· |
| O | 2818 hCoV-19/England/MILK-A7C120/2020\|EPI_ISL_607401\|20 | ···················· | ···················· | ···················· | ···················· | ···················· | ···················· |
| O | 2819 hCoV-19/England/MILK-A7C90A/2020\|EPI_ISL_607402\|20 | ···················· | ···················· | ···················· | ···················· | ···················· | ···················· |
| O | 2820 hCoV-19/England/MILK-A7C5AC/2020\|EPI_ISL_607403\|20 | ···················· | ···················· | ···················· | ···················· | ···················· | ···················· |
| O | 2821 hCoV-19/England/MILK-A7B631/2020\|EPI_ISL_607404\|20 | ···················· | ···················· | ···················· | ···················· | ···················· | ···················· |
| O | 2823 hCoV-19/England/MILK-A7CB9B/2020\|EPI_ISL_607406\|20 | ···················· | ···················· | ···················· | ···················· | ···················· | ···················· |
| O | 2824 hCoV-19/England/MILK-A7B4FB/2020\|EPI_ISL_607407\|20 | ···················· | ···················· | ···················· | ···················· | ···················· | ···················· |
| O | 2825 hCoV-19/England/MILK-A7BD1E/2020\|EPI_ISL_607411\|20 | ···················· | ···················· | ···················· | ···················· | ···················· | ···················· |
| O | 2826 hCoV-19/England/MILK-A7C9A0/2020\|EPI_ISL_607412\|20 | ···················· | ···················· | ···················· | ···················· | ···················· | ···················· |
| O | 2827 hCoV-19/England/MILK-A7B8FF/2020\|EPI_ISL_607413\|20 | ···················· | ···················· | ···················· | ···················· | ···················· | ···················· |
| O | 2828 hCoV-19/England/MILK-A7B77A/2020\|EPI_ISL_607414\|20 | ···················· | ···················· | ···················· | ···················· | ···················· | ···················· |
| O | 2829 hCoV-19/England/MILK-A7B1C7/2020\|EPI_ISL_607415\|20 | ···················· | ·A······A··········· | ···················· | ···················· | ···················· | ···················· |
| O | 2830 hCoV-19/England/MILK-A7C928/2020\|EPI_ISL_607417\|20 | ···················· | ···················· | ···················· | ···················· | ···················· | ···················· |
| O | 2831 hCoV-19/England/MILK-A7C81C/2020\|EPI_ISL_607419\|20 | ···················· | ···················· | ···················· | ···················· | ···················· | ···················· |
| O | 2832 hCoV-19/England/MILK-A7C472/2020\|EPI_ISL_607420\|20 | ···················· | ···················· | ···················· | ···················· | ···················· | ···················· |
| O | 2833 hCoV-19/England/MILK-A7B75C/2020\|EPI_ISL_607421\|20 | ···················· | ···················· | ···················· | ···················· | ···················· | ···················· |
| O | 2834 hCoV-19/England/MILK-A7B4DD/2020\|EPI_ISL_607422\|20 | ···················· | ···················· | ···················· | ···················· | ···················· | ···················· |
| O | 2835 hCoV-19/England/MILK-A7B66E/2020\|EPI_ISL_607424\|20 | ···················· | ···················· | ···················· | ···················· | ···················· | ···················· |
| O | 2836 hCoV-19/England/MILK-A7C7D3/2020\|EPI_ISL_607425\|20 | ···················· | ···················· | ···················· | ···················· | ···················· | ···················· |
| O | 2837 hCoV-19/England/MILK-A7CC01/2020\|EPI_ISL_607427\|20 | ···················· | ···················· | ···················· | ···················· | ···················· | ···················· |
| O | 2838 hCoV-19/England/MILK-A7B76B/2020\|EPI_ISL_607429\|20 | ···················· | ···················· | ···················· | ···················· | ···················· | ···················· |
| O | 2839 hCoV-19/England/MILK-A6BC32/2020\|EPI_ISL_608721\|20 | ···················· | ···················· | ···················· | ···················· | ···················· | ···················· |
| O | 2840 hCoV-19/England/MILK-A6B458/2020\|EPI_ISL_608722\|20 | ···················· | ·A·················· | ···················· | ···················· | ···················· | ···A················ |
| O | 2841 hCoV-19/England/MILK-A6B616/2020\|EPI_ISL_608723\|20 | ···················· | ···················· | ···················· | ···················· | ···················· | ···················· |
| O | 2842 hCoV-19/England/MILK-A69F0E/2020\|EPI_ISL_608725\|20 | ···················· | ···················· | ···················· | ···················· | ···················· | ···················· |
| O | 2843 hCoV-19/England/MILK-A69D13/2020\|EPI_ISL_608726\|20 | ···················· | ···················· | ···················· | ···················· | ···················· | ···················· |
| O | 2844 hCoV-19/England/MILK-A69FE0/2020\|EPI_ISL_608732\|20 | ···················· | ···················· | ···················· | ···················· | ···················· | ···················· |
| O | 2845 hCoV-19/England/MILK-A6B28B/2020\|EPI_ISL_608733\|20 | ···················· | ···················· | ···················· | ···················· | ···················· | ···················· |
| O | 2846 hCoV-19/England/MILK-A6BAB0/2020\|EPI_ISL_608734\|20 | ···················· | ···················· | ···················· | ···················· | ···················· | ···················· |
| O | 2847 hCoV-19/England/MILK-A6B555/2020\|EPI_ISL_608735\|20 | ···················· | ···················· | ···················· | ···················· | ···················· | ···················· |
| O | 2848 hCoV-19/England/MILK-A6B29A/2020\|EPI_ISL_608737\|20 | ···················· | ···················· | ···················· | ···················· | ···················· | ···················· |
| O | 2849 hCoV-19/England/MILK-A6A18F/2020\|EPI_ISL_608738\|20 | ···················· | ···················· | ···················· | ···················· | ···················· | ···················· |
| O | 2850 hCoV-19/England/MILK-A6AD30/2020\|EPI_ISL_608739\|20 | ···················· | ···················· | ···················· | ···················· | ···················· | ···················· |
| O | 2851 hCoV-19/England/MILK-A69CDA/2020\|EPI_ISL_608740\|20 | ···················· | ···················· | ···················· | ···················· | ···················· | ···················· |
| O | 2852 hCoV-19/England/MILK-A69C70/2020\|EPI_ISL_608742\|20 | ···················· | ···················· | ···················· | ···················· | ···················· | ···················· |
| O | 2853 hCoV-19/England/MILK-A6BACF/2020\|EPI_ISL_608743\|20 | ···················· | ···················· | ···················· | ···················· | ···················· | ···················· |
| O | 2854 hCoV-19/England/MILK-A6B2E5/2020\|EPI_ISL_608744\|20 | ···················· | ···················· | ···················· | ···················· | ···················· | ···················· |
| O | 2855 hCoV-19/England/MILK-A6B1AC/2020\|EPI_ISL_608745\|20 | ···················· | ···················· | ···················· | ···················· | ···················· | ···················· |
| O | 2856 hCoV-19/England/MILK-A69BA0/2020\|EPI_ISL_608746\|20 | ···················· | ···················· | ···················· | ···················· | ···················· | ···················· |
| O | 2857 hCoV-19/England/MILK-A6AF67/2020\|EPI_ISL_608747\|20 | ···················· | ·A·················· | ···················· | ···················· | ···················· | ···················· |
| O | 2858 hCoV-19/England/MILK-A6AF94/2020\|EPI_ISL_608749\|20 | ···················· | ·A·················· | ···················· | ···················· | ···················· | ···A················ |
| O | 2859 hCoV-19/England/MILK-A6AC51/2020\|EPI_ISL_608750\|20 | ···················· | ···················· | ···················· | ···················· | ···················· | ···················· |
| O | 2860 hCoV-19/England/MILK-A69EC5/2020\|EPI_ISL_608751\|20 | ···················· | ···················· | ···················· | ···················· | ···················· | ···················· |
| O | 2861 hCoV-19/England/MILK-A69E3E/2020\|EPI_ISL_608753\|20 | ···················· | ···················· | ···················· | ···················· | ···················· | ···················· |
| O | 2862 hCoV-19/England/MILK-A6BA74/2020\|EPI_ISL_608755\|20 | ···················· | ···················· | ···················· | ···················· | ···················· | ···················· |
| O | 2863 hCoV-19/England/MILK-A69C25/2020\|EPI_ISL_608756\|20 | ···················· | ···················· | ···················· | ···················· | ···················· | ···················· |
| O | 2864 hCoV-19/England/MILK-A6B4A3/2020\|EPI_ISL_608757\|20 | ···················· | ···················· | ···················· | ···················· | ···················· | ···················· |
| O | 2865 hCoV-19/England/MILK-A69DE6/2020\|EPI_ISL_608758\|20 | ···················· | ·A·················· | ···················· | ···················· | ···················· | ···A················ |
| O | 2866 hCoV-19/Wales/ALDP-A4FF2C/2020\|EPI_ISL_609210\|2020 | ···················· | ···················· | ···················· | ···················· | ···················· | ···················· |
| O | 2867 hCoV-19/Wales/ALDP-A4FF1D/2020\|EPI_ISL_609211\|2020 | ···················· | ···················· | ···················· | ···················· | ·····T·············· | ···················· |
| O | 2868 hCoV-19/Wales/ALDP-A508E1/2020\|EPI_ISL_609212\|2020 | ···················· | ···················· | ···················· | ···················· | ···················· | ···················· |
| O | 2869 hCoV-19/Wales/ALDP-A4FEE3/2020\|EPI_ISL_609213\|2020 | ···················· | ···················· | ···················· | ···················· | ···················· | ···················· |
| O | 2870 hCoV-19/Wales/ALDP-A51AF9/2020\|EPI_ISL_609215\|2020 | ···················· | ···················· | ···················· | ···················· | ···················· | ···················· |
| O | 2871 hCoV-19/Wales/ALDP-A52982/2020\|EPI_ISL_609216\|2020 | ···················· | ···················· | ···················· | ···················· | ···················· | ···················· |
| O | 2872 hCoV-19/Wales/ALDP-A51B9C/2020\|EPI_ISL_609217\|2020 | ···················· | ···················· | ···················· | ···················· | ···················· | ···················· |
| O | 2873 hCoV-19/Wales/ALDP-A52991/2020\|EPI_ISL_609218\|2020 | ···················· | ···················· | ···················· | ···················· | ···················· | ···················· |
| O | 2874 hCoV-19/Wales/MILK-A46BE5/2020\|EPI_ISL_609220\|2020 | ···················· | ···················· | ···················· | ···················· | ···················· | ···················· |
| O | 2875 hCoV-19/Wales/MILK-A47E63/2020\|EPI_ISL_609221\|2020 | ···················· | ···················· | ···················· | ···················· | ···················· | ···················· |
| O | 2876 hCoV-19/Wales/MILK-A46C00/2020\|EPI_ISL_609222\|2020 | ···················· | ···················· | ···················· | ···················· | ···················· | ···················· |
| O | 2877 hCoV-19/Wales/MILK-A47C4B/2020\|EPI_ISL_609223\|2020 | ···················· | ···················· | ···················· | ···················· | ···················· | ···················· |
| O | 2878 hCoV-19/Wales/MILK-A473FB/2020\|EPI_ISL_609225\|2020 | ···················· | ···················· | ···················· | ···················· | ···················· | ···················· |
| O | 2879 hCoV-19/Wales/MILK-A47A50/2020\|EPI_ISL_609226\|2020 | ···················· | ···················· | ···················· | ···················· | ···················· | ···················· |
| O | 2880 hCoV-19/Wales/MILK-A46BF4/2020\|EPI_ISL_609229\|2020 | ···················· | ···················· | ···················· | ···················· | ···················· | ···················· |
| O | 2881 hCoV-19/Wales/MILK-A47D84/2020\|EPI_ISL_609231\|2020 | ···················· | ···················· | ···················· | ···················· | ···················· | ···················· |
| O | 2882 hCoV-19/Wales/MILK-A46C2E/2020\|EPI_ISL_609232\|2020 | ···················· | ···················· | ···················· | ···················· | ···················· | ···················· |
| O | 2883 hCoV-19/Wales/MILK-A47470/2020\|EPI_ISL_609233\|2020 | ···················· | ···················· | ···················· | ···················· | ···················· | ···················· |
| O | 2884 hCoV-19/Wales/MILK-A46B9A/2020\|EPI_ISL_609234\|2020 | ···················· | ···················· | ···················· | ···················· | ···················· | ···················· |

O 2885 hCoV-19/Wales/MILK-A473CE/2020|EPI_ISL_609235|2020 .................... .................... .................... .................... .................... ....................
O 2886 hCoV-19/Wales/MILK-A477FF/2020|EPI_ISL_609236|2020 .................... .................... .................... .................... .................... ....................
O 2887 hCoV-19/Wales/MILK-A474AD/2020|EPI_ISL_609238|2020 .................... .................... .................... .................... .................... ....................
O 2888 hCoV-19/Wales/MILK-A46E46/2020|EPI_ISL_609239|2020 .................... .................... .................... .................... .................... ....................
O 2889 hCoV-19/Wales/MILK-A47777/2020|EPI_ISL_609241|2020 .................... .................... .................... .................... .................... ....................
O 2890 hCoV-19/Wales/MILK-A47829/2020|EPI_ISL_609242|2020 .................... .................... .................... .................... .................... ....................
O 2891 hCoV-19/Wales/MILK-A47AAB/2020|EPI_ISL_609243|2020 .................... .................... .................... .................... .................... ....................
O 2892 hCoV-19/Wales/MILK-A477D1/2020|EPI_ISL_609244|2020 .................... .................... .................... .................... .................... ....................
O 2893 hCoV-19/Wales/MILK-A47C2D/2020|EPI_ISL_609245|2020 .................... .................... .................... .................... .................... ....................
O 2894 hCoV-19/Wales/MILK-A46DB2/2020|EPI_ISL_609246|2020 .................... .................... .................... .................... .................... ....................
O 2895 hCoV-19/Wales/MILK-A4772C/2020|EPI_ISL_609247|2020 .................... .................... .................... .................... .................... ....................
O 2896 hCoV-19/Wales/MILK-A4774A/2020|EPI_ISL_609248|2020 .................... .................... .................... .................... .................... ....................
O 2897 hCoV-19/Wales/MILK-A46990/2020|EPI_ISL_609249|2020 .................... .................... .................... .................... .................... ....................
O 2898 hCoV-19/Wales/MILK-A4771D/2020|EPI_ISL_609270|2020 .................... .................... .................... .................... .................... ....................
O 2899 hCoV-19/Wales/MILK-A46E19/2020|EPI_ISL_609271|2020 .................... .................... .................... .................... .................... ....................
O 2900 hCoV-19/England/MILK-A4803F/2020|EPI_ISL_609272|20 .................... .................... .................... .................... .................... ....................
O 2901 hCoV-19/Wales/MILK-A47D66/2020|EPI_ISL_609273|2020 .................... .................... .................... .................... .................... ....................
O 2902 hCoV-19/Wales/MILK-A46BB8/2020|EPI_ISL_609276|2020 .................... .................... .................... .................... .................... ....................
O 2903 hCoV-19/Wales/MILK-A47759/2020|EPI_ISL_609277|2020 .................... .................... .................... .................... .................... ....................
O 2904 hCoV-19/Wales/MILK-A47A7E/2020|EPI_ISL_609278|2020 .................... .................... .................... .................... .................... ....................
O 2905 hCoV-19/Wales/MILK-A47B4E/2020|EPI_ISL_609279|2020 .................... .................... .................... .................... .................... ....................
O 2906 hCoV-19/Wales/MILK-A47EDC/2020|EPI_ISL_609280|2020 .................... .................... .................... .................... .................... ....................
O 2907 hCoV-19/Wales/MILK-A4680C/2020|EPI_ISL_609281|2020 .................... .................... .................... .................... .................... ....................
O 2908 hCoV-19/Wales/MILK-A463DE/2020|EPI_ISL_609282|2020 .................... .................... .................... .................... .................... ....................
O 2909 hCoV-19/Wales/MILK-A46884/2020|EPI_ISL_609283|2020 .................... .................... .................... .................... .................... ....................
O 2910 hCoV-19/Wales/MILK-A46356/2020|EPI_ISL_609284|2020 .................... .................... .................... .................... .................... ....................
O 2911 hCoV-19/Wales/MILK-A46374/2020|EPI_ISL_609288|2020 .................... .................... .................... .................... .................... ....................
O 2912 hCoV-19/Russia/CRIE4907775/2020|EPI_ISL_610227|202 .................... .................... .................... .................... .................... ....................
O 2913 hCoV-19/Russia/CRIE4978885/2020|EPI_ISL_610228|202 .................... .................... .................... .................... .................... ....................
O 2914 hCoV-19/Russia/CRIE4973936/2020|EPI_ISL_610229|202 .................... .................... .................... .................... .................... ....................
O 2915 hCoV-19/Russia/CRIE5030221/2020|EPI_ISL_610230|202 .................... .................... .................... .................... .................... ....................
O 2916 hCoV-19/Russia/CRIE5031464/2020|EPI_ISL_610231|202 .................... .................... .................... .................... .................... ....................
O 2917 hCoV-19/Russia/CRIE4991529/2020|EPI_ISL_610232|202 .................... .................... .................... .................... .................... ....................
O 2918 hCoV-19/Russia/CRIE4990266/2020|EPI_ISL_610233|202 .................... .................... .................... .................... .................... ....................
O 2919 hCoV-19/Russia/CRIE4988935/2020|EPI_ISL_610234|202 ...T................ .................... .................... .................... .................... ....................
O 2920 hCoV-19/Russia/CRIE5021376/2020|EPI_ISL_610235|202 .................... .................... .................... .................... .................... ....................
O 2921 hCoV-19/Russia/CRIE5047165/2020|EPI_ISL_610236|202 .................... .................... .................... .................... .................... ....................
O 2922 hCoV-19/Russia/CRIE170668/2020|EPI_ISL_610237|2020 .................... .................... .................... .................... .................... ....................
O 2923 hCoV-19/USA/NM-UNM-00794/2020|EPI_ISL_610260|2020- .................... .................... .................... .................... .................... ....................
O 2924 hCoV-19/USA/NM-UNM-00797/2020|EPI_ISL_610261|2020- .................... .................... .................... .................... .................... ....................
O 2925 hCoV-19/USA/NM-UNM-00799/2020|EPI_ISL_610262|2020- .................... .................... .................... .................... .................... ....................
O 2926 hCoV-19/USA/NM-UNM-00815/2020|EPI_ISL_610265|2020- .................... .................... .................... .................... .................... ....................
O 2927 hCoV-19/USA/NM-UNM-00818/2020|EPI_ISL_610266|2020- .................... .................... .................... .................... .................... ....................
O 2928 hCoV-19/USA/NM-UNM-00819/2020|EPI_ISL_610267|2020- .................... .................... .................... .................... .................... ....................
O 2929 hCoV-19/USA/NM-UNM-00830/2020|EPI_ISL_610268|2020- .................... .................... .................... .................... .................... ....................
O 2930 hCoV-19/USA/NM-UNM-00832/2020|EPI_ISL_610269|2020- .................... .................... .................... .................... .................... ....................
O 2931 hCoV-19/USA/NM-UNM-00833/2020|EPI_ISL_610270|2020- .................... .................... .................... .................... .................... ....................
O 2932 hCoV-19/USA/NM-UNM-00834/2020|EPI_ISL_610271|2020- .................... .................... .................... .................... .................... ....................
O 2933 hCoV-19/USA/NM-UNM-00838/2020|EPI_ISL_610272|2020- .................... .................... .................... .................... .................... ....................
O 2934 hCoV-19/USA/NM-UNM-00840/2020|EPI_ISL_610273|2020- .................... .................... .................... .................... .................... ....................
O 2935 hCoV-19/USA/NM-UNM-00841/2020|EPI_ISL_610274|2020- .................... .................... .................... .................... .................... ....................
O 2936 hCoV-19/USA/NM-UNM-00845/2020|EPI_ISL_610275|2020- .................... .................... .................... .................... .................... ....................
O 2937 hCoV-19/USA/NM-UNM-00846/2020|EPI_ISL_610276|2020- .................... .................... .................... .................... .................... ....................
O 2938 hCoV-19/USA/NM-UNM-00849/2020|EPI_ISL_610277|2020- .................... .................... .................... .................... .................... ....................
O 2939 hCoV-19/USA/NM-UNM-00850/2020|EPI_ISL_610278|2020- .................... .................... .................... .................... .................... ....................
O 2940 hCoV-19/USA/NM-UNM-00851/2020|EPI_ISL_610279|2020- .................... .................... .................... .................... .................... ....................
O 2941 hCoV-19/Wales/PHWC-48016B/2020|EPI_ISL_612856|2020 .................... .................... .................... .................... .................... ....................
O 2942 hCoV-19/Wales/PHWC-48017A/2020|EPI_ISL_612857|2020 .................... .................... .................... .................... .................... ....................
O 2943 hCoV-19/Wales/PHWC-480189/2020|EPI_ISL_612858|2020 .................... .................... .................... .................... .................... ....................
O 2944 hCoV-19/Wales/PHWC-480198/2020|EPI_ISL_612859|2020 .................... .................... .................... .................... .................... ....................
O 2945 hCoV-19/Wales/PHWC-4801B6/2020|EPI_ISL_612860|2020 .................... .................... .................... .................... .................... ....................
O 2946 hCoV-19/Wales/PHWC-4801D4/2020|EPI_ISL_612861|2020 .................... .................... .................... .................... .................... ....................
O 2947 hCoV-19/Wales/PHWC-4801E3/2020|EPI_ISL_612862|2020 .................... .................... .................... .................... .................... ....................
O 2948 hCoV-19/Wales/PHWC-48020E/2020|EPI_ISL_612863|2020 .................... .................... .................... .................... .................... ....................
O 2949 hCoV-19/Wales/PHWC-48021D/2020|EPI_ISL_612864|2020 .................... .................... .................... .................... .................... ....................
O 2950 hCoV-19/Wales/PHWC-48022C/2020|EPI_ISL_612865|2020 .................... .................... .................... .................... .................... ....................
O 2951 hCoV-19/Wales/PHWC-48023B/2020|EPI_ISL_612866|2020 .................... .................... .................... .................... .................... ....................
O 2954 hCoV-19/Wales/PHWC-480286/2020|EPI_ISL_612869|2020 .................... .................... .................... .................... .................... ....................
O 2955 hCoV-19/Wales/PHWC-4802A4/2020|EPI_ISL_612870|2020 .................... .................... .................... .................... .................... ....................
O 2956 hCoV-19/Wales/PHWC-4802C2/2020|EPI_ISL_612871|2020 .................... .................... .................... .................... .................... ....................
O 2957 hCoV-19/Wales/PHWC-4802D1/2020|EPI_ISL_612872|2020 .................... .................... .................... .................... .................... ....................
O 2958 hCoV-19/Wales/PHWC-48030B/2020|EPI_ISL_612873|2020 .................... .................... .................... .................... .................... ....................
O 2959 hCoV-19/Wales/PHWC-480329/2020|EPI_ISL_612874|2020 .................... .................... .................... .................... .................... ....................
O 2960 hCoV-19/Wales/PHWC-480338/2020|EPI_ISL_612875|2020 .................... .................... .................... .................... .................... ....................
O 2961 hCoV-19/Wales/PHWC-480347/2020|EPI_ISL_612876|2020 .................... .................... .................... .................... .................... ....................
O 2962 hCoV-19/Wales/PHWC-480374/2020|EPI_ISL_612877|2020 .................... .................... .................... .................... .................... ....................
O 2963 hCoV-19/Wales/PHWC-480383/2020|EPI_ISL_612878|2020 .................... .................... .................... .................... .................... ....................
O 2964 hCoV-19/Wales/PHWC-480392/2020|EPI_ISL_612879|2020 .................... .................... .................... .................... .................... ....................
O 2965 hCoV-19/Wales/PHWC-4803A1/2020|EPI_ISL_612880|2020 .................... .................... .................... .................... .................... ....................
O 2966 hCoV-19/Wales/PHWC-4803B0/2020|EPI_ISL_612881|2020 .................... .................... .................... .................... .................... ....................

| | | | | | | | | |
|---|---|---|---|---|---|---|---|---|
| O | 2967 | hCoV-19/Wales/PHWC-4803CF/2020\|EPI_ISL_612882\|2020 | .................... | .................... | .................... | .................... | .................... | .................... |
| O | 2968 | hCoV-19/Wales/PHWC-4803FC/2020\|EPI_ISL_612883\|2020 | .................... | .................... | .................... | .................... | .................... | .................... |
| O | 2969 | hCoV-19/Wales/PHWC-480408/2020\|EPI_ISL_612884\|2020 | .................... | .................... | .................... | .................... | .................... | .................... |
| O | 2970 | hCoV-19/Wales/PHWC-480417/2020\|EPI_ISL_612885\|2020 | .................... | .................... | .................... | .................... | .................... | .................... |
| O | 2971 | hCoV-19/Wales/PHWC-480426/2020\|EPI_ISL_612886\|2020 | .................... | .................... | .................... | .................... | .................... | .................... |
| O | 2972 | hCoV-19/Wales/PHWC-480435/2020\|EPI_ISL_612887\|2020 | .................... | .................... | .................... | .................... | .................... | .................... |
| O | 2973 | hCoV-19/Wales/PHWC-480471/2020\|EPI_ISL_612888\|2020 | .................... | .................... | .................... | .................... | .................... | .................... |
| O | 2974 | hCoV-19/Wales/PHWC-480480/2020\|EPI_ISL_612889\|2020 | .................... | .................... | .................... | .................... | .................... | .................... |
| O | 2975 | hCoV-19/Wales/PHWC-48049F/2020\|EPI_ISL_612890\|2020 | .................... | .................... | .................... | .................... | .................... | .................... |
| O | 2976 | hCoV-19/Wales/PHWC-4804AE/2020\|EPI_ISL_612891\|2020 | .................... | .................... | .................... | .................... | .................... | .................... |
| O | 2977 | hCoV-19/Wales/PHWC-4804BD/2020\|EPI_ISL_612892\|2020 | .................... | .................... | .................... | .................... | .................... | .................... |
| O | 2978 | hCoV-19/Wales/PHWC-4804DB/2020\|EPI_ISL_612893\|2020 | .................... | .................... | .................... | .................... | .................... | .................... |
| O | 2979 | hCoV-19/Wales/PHWC-4804EA/2020\|EPI_ISL_612894\|2020 | .................... | .................... | .................... | .................... | .................... | .................... |
| O | 2980 | hCoV-19/Wales/PHWC-4804F9/2020\|EPI_ISL_612895\|2020 | .................... | .................... | .................... | .................... | .................... | .................... |
| O | 2981 | hCoV-19/Wales/PHWC-480505/2020\|EPI_ISL_612896\|2020 | .................... | .................... | .................... | .................... | .................... | .................... |
| O | 2982 | hCoV-19/Wales/PHWC-480514/2020\|EPI_ISL_612897\|2020 | .................... | .................... | .................... | .................... | .................... | .................... |
| O | 2983 | hCoV-19/Wales/PHWC-480523/2020\|EPI_ISL_612898\|2020 | .................... | .................... | .................... | .................... | .................... | .................... |
| O | 2984 | hCoV-19/Wales/PHWC-480532/2020\|EPI_ISL_612899\|2020 | .................... | .................... | .................... | .................... | .................... | .................... |
| O | 2986 | hCoV-19/Wales/PHWC-48214B/2020\|EPI_ISL_613201\|2020 | .................... | .................... | .................... | .................... | .................... | .................... |
| O | 2987 | hCoV-19/Wales/PHWC-48215A/2020\|EPI_ISL_613202\|2020 | .................... | .................... | .................... | .................... | .................... | .................... |
| O | 2988 | hCoV-19/Wales/PHWC-482196/2020\|EPI_ISL_613203\|2020 | .................... | .................... | .................... | .................... | .................... | .................... |
| O | 2989 | hCoV-19/Wales/PHWC-48220C/2020\|EPI_ISL_613204\|2020 | .................... | .................... | .................... | .................... | .................... | .................... |
| O | 2990 | hCoV-19/Wales/PHWC-48221B/2020\|EPI_ISL_613205\|2020 | .................... | .................... | .................... | .................... | .................... | .................... |
| O | 2991 | hCoV-19/Wales/PHWC-482239/2020\|EPI_ISL_613206\|2020 | .................... | .................... | .................... | .................... | .................... | .................... |
| O | 2992 | hCoV-19/Wales/PHWC-482257/2020\|EPI_ISL_613207\|2020 | .................... | .................... | .................... | .................... | .................... | .................... |
| O | 2993 | hCoV-19/Wales/PHWC-482284/2020\|EPI_ISL_613208\|2020 | .................... | .................... | .................... | .................... | .................... | .................... |
| O | 2994 | hCoV-19/Wales/PHWC-482293/2020\|EPI_ISL_613209\|2020 | .................... | .................... | .................... | .................... | .................... | .................... |
| O | 2995 | hCoV-19/Wales/PHWC-4822A2/2020\|EPI_ISL_613210\|2020 | .................... | .................... | .................... | .................... | .................... | .................... |
| O | 2996 | hCoV-19/Wales/PHWC-4822FD/2020\|EPI_ISL_613211\|2020 | .................... | .................... | .................... | .................... | .................... | .................... |
| O | 2997 | hCoV-19/Wales/PHWC-482309/2020\|EPI_ISL_613212\|2020 | .................... | .................... | .................... | .................... | .................... | .................... |
| O | 2998 | hCoV-19/Wales/PHWC-482336/2020\|EPI_ISL_613213\|2020 | .................... | .................... | .................... | .................... | .................... | .................... |
| O | 2999 | hCoV-19/Wales/PHWC-482354/2020\|EPI_ISL_613214\|2020 | .................... | .................... | .................... | .................... | .................... | .................... |
| O | 3000 | hCoV-19/Wales/PHWC-482390/2020\|EPI_ISL_613215\|2020 | .................... | .................... | .................... | .................... | .................... | .................... |
| O | 3001 | hCoV-19/Wales/PHWC-4823AF/2020\|EPI_ISL_613216\|2020 | .................... | .................... | .................... | .................... | .................... | .................... |
| O | 3002 | hCoV-19/Wales/PHWC-4823BE/2020\|EPI_ISL_613217\|2020 | .................... | .................... | .................... | .................... | .................... | .................... |
| O | 3003 | hCoV-19/Wales/PHWC-4823FA/2020\|EPI_ISL_613218\|2020 | .................... | .................... | .................... | .................... | .................... | .................... |
| O | 3004 | hCoV-19/Wales/PHWC-482433/2020\|EPI_ISL_613219\|2020 | .................... | .................... | .................... | .................... | .................... | .................... |
| O | 3005 | hCoV-19/Wales/PHWC-482460/2020\|EPI_ISL_613220\|2020 | .................... | .................... | .................... | .................... | .................... | .................... |
| O | 3006 | hCoV-19/Wales/PHWC-48247F/2020\|EPI_ISL_613221\|2020 | .................... | .................... | .................... | .................... | .................... | .................... |
| O | 3007 | hCoV-19/Wales/PHWC-48248E/2020\|EPI_ISL_613222\|2020 | .................... | .................... | .................... | .................... | .................... | .................... |
| O | 3008 | hCoV-19/Wales/PHWC-4824AC/2020\|EPI_ISL_613223\|2020 | .................... | .................... | .................... | .................... | .................... | .................... |
| O | 3009 | hCoV-19/Wales/PHWC-4824CA/2020\|EPI_ISL_613224\|2020 | .................... | .................... | .................... | .................... | .................... | .................... |
| O | 3010 | hCoV-19/Wales/PHWC-4824E8/2020\|EPI_ISL_613225\|2020 | .................... | .................... | .................... | .................... | .................... | .................... |
| O | 3011 | hCoV-19/Wales/PHWC-482521/2020\|EPI_ISL_613226\|2020 | .................... | .................... | .................... | .................... | .................... | .................... |
| O | 3012 | hCoV-19/Wales/PHWC-48254F/2020\|EPI_ISL_613227\|2020 | .................... | .................... | .................... | .................... | .................... | .................... |
| O | 3013 | hCoV-19/Wales/PHWC-48255E/2020\|EPI_ISL_613228\|2020 | .................... | .................... | .................... | .................... | .................... | .................... |
| O | 3014 | hCoV-19/Wales/PHWC-48256D/2020\|EPI_ISL_613229\|2020 | .................... | .................... | .................... | .................... | .................... | .................... |
| O | 3015 | hCoV-19/Wales/PHWC-4825A9/2020\|EPI_ISL_613230\|2020 | .................... | .................... | .................... | .................... | .................... | .................... |
| O | 3016 | hCoV-19/Wales/PHWC-4825D6/2020\|EPI_ISL_613231\|2020 | .................... | .................... | .................... | .................... | .................... | .................... |
| O | 3017 | hCoV-19/Wales/PHWC-482600/2020\|EPI_ISL_613232\|2020 | .................... | .................... | .................... | .................... | .................... | .................... |
| O | 3019 | hCoV-19/Wales/PHWC-48264C/2020\|EPI_ISL_613234\|2020 | .................... | .................... | .................... | .................... | .................... | .................... |
| O | 3021 | hCoV-19/Wales/PHWC-4826C4/2020\|EPI_ISL_613236\|2020 | .................... | .................... | .................... | .................... | .................... | .................... |
| O | 3022 | hCoV-19/Wales/PHWC-4826D3/2020\|EPI_ISL_613237\|2020 | .................... | .................... | .................... | .................... | .................... | .................... |
| O | 3023 | hCoV-19/Wales/PHWC-4826E2/2020\|EPI_ISL_613238\|2020 | .................... | .................... | .................... | .................... | .................... | .................... |
| O | 3024 | hCoV-19/Wales/PHWC-48272B/2020\|EPI_ISL_613239\|2020 | .................... | .................... | .................... | .................... | .................... | .................... |
| O | 3025 | hCoV-19/Wales/PHWC-482785/2020\|EPI_ISL_613240\|2020 | .................... | .................... | .................... | .................... | .................... | .................... |
| O | 3026 | hCoV-19/Wales/PHWC-482794/2020\|EPI_ISL_613241\|2020 | .................... | .................... | .................... | .................... | .................... | .................... |
| O | 3028 | hCoV-19/Wales/PHWC-4827C1/2020\|EPI_ISL_613243\|2020 | .................... | .................... | .................... | .................... | .................... | .................... |
| O | 3029 | hCoV-19/Wales/PHWC-4827D0/2020\|EPI_ISL_613244\|2020 | .................... | .................... | .................... | .................... | .................... | .................... |
| O | 3030 | hCoV-19/Wales/PHWC-4827FE/2020\|EPI_ISL_613245\|2020 | .................... | .................... | .................... | .................... | .................... | .................... |
| O | 3031 | hCoV-19/Wales/PHWC-482819/2020\|EPI_ISL_613246\|2020 | .................... | .................... | .................... | .................... | .................... | .................... |
| O | 3032 | hCoV-19/Wales/PHWC-482828/2020\|EPI_ISL_613247\|2020 | .................... | .................... | .................... | .................... | .................... | .................... |
| O | 3033 | hCoV-19/Wales/PHWC-482864/2020\|EPI_ISL_613248\|2020 | .................... | .................... | .................... | .................... | .................... | .................... |
| O | 3034 | hCoV-19/Wales/PHWC-482873/2020\|EPI_ISL_613249\|2020 | .................... | .................... | .................... | .................... | .................... | .................... |
| O | 3037 | hCoV-19/Wales/PHWC-482907/2020\|EPI_ISL_613252\|2020 | .................... | .................... | .................... | .................... | .................... | .................... |
| O | 3038 | hCoV-19/Wales/PHWC-482934/2020\|EPI_ISL_613253\|2020 | .................... | .................... | .................... | .................... | .................... | .................... |
| O | 3040 | hCoV-19/Wales/PHWC-482961/2020\|EPI_ISL_613255\|2020 | .................... | .................... | .................... | .................... | .................... | .................... |
| O | 3042 | hCoV-19/Wales/PHWC-4829DA/2020\|EPI_ISL_613257\|2020 | .................... | .................... | .................... | .................... | .................... | .................... |
| O | 3043 | hCoV-19/Wales/PHWC-4829E9/2020\|EPI_ISL_613258\|2020 | .................... | .................... | .................... | .................... | .................... | .................... |
| O | 3044 | hCoV-19/Wales/PHWC-482A04/2020\|EPI_ISL_613259\|2020 | .................... | .................... | .................... | .................... | .................... | .................... |
| O | 3045 | hCoV-19/Wales/PHWC-482A13/2020\|EPI_ISL_613260\|2020 | .................... | .................... | .................... | .................... | .................... | .................... |
| O | 3046 | hCoV-19/Wales/PHWC-482A31/2020\|EPI_ISL_613261\|2020 | .................... | .................... | .................... | .................... | .................... | .................... |
| O | 3047 | hCoV-19/Wales/PHWC-482A40/2020\|EPI_ISL_613262\|2020 | .................... | .................... | .................... | .................... | .................... | .................... |
| O | 3048 | hCoV-19/Wales/PHWC-482A8C/2020\|EPI_ISL_613263\|2020 | .................... | .................... | .................... | .................... | .................... | .................... |
| O | 3049 | hCoV-19/Wales/PHWC-482AAA/2020\|EPI_ISL_613264\|2020 | .................... | .................... | .................... | .................... | .................... | .................... |
| O | 3050 | hCoV-19/Wales/PHWC-482AC8/2020\|EPI_ISL_613265\|2020 | .................... | .................... | .................... | .................... | .................... | .................... |
| O | 3051 | hCoV-19/Wales/PHWC-482AD7/2020\|EPI_ISL_613266\|2020 | .................... | .................... | .................... | .................... | .................... | .................... |
| O | 3052 | hCoV-19/Wales/PHWC-482B3E/2020\|EPI_ISL_613267\|2020 | .................... | .................... | .................... | .................... | .................... | .................... |
| O | 3053 | hCoV-19/Wales/PHWC-482B5C/2020\|EPI_ISL_613268\|2020 | .................... | .................... | .................... | .................... | .................... | .................... |
| O | 3054 | hCoV-19/Wales/PHWC-482B6B/2020\|EPI_ISL_613269\|2020 | .................... | .................... | .................... | .................... | .................... | .................... |

O 3055 hCoV-19/Wales/PHWC-482B98/2020|EPI_ISL_613270|2020 ···················· ···················· ···················· ···················· ···················· ····················
O 3056 hCoV-19/Wales/PHWC-482BA7/2020|EPI_ISL_613271|2020 ···················· ···················· ···················· ···················· ···················· ····················
O 3057 hCoV-19/Wales/PHWC-482BB6/2020|EPI_ISL_613272|2020 ···················· ···················· ···················· ···················· ···················· ····················
O 3058 hCoV-19/Wales/PHWC-482BE3/2020|EPI_ISL_613273|2020 ···················· ···················· ···················· ···················· ···················· ····················
O 3059 hCoV-19/Wales/PHWC-482C0E/2020|EPI_ISL_613274|2020 ···················· ···················· ···················· ···················· ···················· ····················
O 3060 hCoV-19/Wales/PHWC-482C2C/2020|EPI_ISL_613275|2020 ···················· ···················· ···················· ···················· ···················· ····················
O 3061 hCoV-19/Wales/PHWC-482C3B/2020|EPI_ISL_613276|2020 ···················· ···················· ···················· ···················· ···················· ····················
O 3062 hCoV-19/Wales/PHWC-482C86/2020|EPI_ISL_613277|2020 ···················· ·A·················· ···················· ···················· ···················· ··A·················
O 3063 hCoV-19/Wales/PHWC-482CA4/2020|EPI_ISL_613278|2020 ···················· ···················· ···················· ···················· ···················· ····················
O 3064 hCoV-19/Wales/PHWC-482CD1/2020|EPI_ISL_613279|2020 ···················· ···················· ···················· ···················· ···················· ····················
O 3065 hCoV-19/Wales/PHWC-482CE0/2020|EPI_ISL_613280|2020 ···················· ···················· ···················· ···················· N··················· ····················
O 3066 hCoV-19/Wales/PHWC-482D1A/2020|EPI_ISL_613281|2020 ···················· ···················· ···················· ···················· ···················· ····················
O 3067 hCoV-19/England/PORT-2D213F/2020|EPI_ISL_613282|20 ···················· ···················· ···················· ···················· ···················· ····················
O 3068 hCoV-19/England/PORT-2D21A8/2020|EPI_ISL_613283|20 ···················· ···················· ···················· ···················· ···················· ····················
O 3069 hCoV-19/England/PORT-2D2287/2020|EPI_ISL_613286|20 ···················· ···················· ···················· ···················· ···················· ····················
O 3070 hCoV-19/England/PORT-2D22E1/2020|EPI_ISL_613287|20 ···················· ···················· ···················· ···················· ···················· ····················
O 3071 hCoV-19/England/PORT-2D465C/2020|EPI_ISL_613288|20 ···················· ···················· ···················· ···················· ···················· ····················
O 3072 hCoV-19/England/PORT-2D4698/2020|EPI_ISL_613289|20 ···················· ···················· ···················· ···················· ·····T·············· ····················
O 3073 hCoV-19/England/ALDP-A861AB/2020|EPI_ISL_606000|20 ···················· ···················· ···················· ···················· ···················· ····················
O 3074 hCoV-19/England/ALDP-A86572/2020|EPI_ISL_606002|20 ···················· ···················· ···················· ···················· ···················· ····················
O 3075 hCoV-19/England/ALDP-A86F38/2020|EPI_ISL_606004|20 ···················· ···················· ···················· ···················· ···················· ····················
O 3076 hCoV-19/England/ALDP-A86DA7/2020|EPI_ISL_606005|20 ···················· ···················· ···················· ···················· ···················· ····················
O 3077 hCoV-19/England/ALDP-A87993/2020|EPI_ISL_606006|20 ···················· ···················· ···················· ···················· ···················· ····················
O 3078 hCoV-19/England/ALDP-A86D01/2020|EPI_ISL_606007|20 ···················· ···················· ···················· ···················· ···················· ····················
O 3079 hCoV-19/England/ALDP-A86C5F/2020|EPI_ISL_606008|20 ···················· ···················· ···················· ···················· ···················· ····················
O 3080 hCoV-19/England/ALDP-A8784B/2020|EPI_ISL_606009|20 ···················· ···················· ···················· ···················· ···················· ····················
O 3081 hCoV-19/England/ALDP-A873C2/2020|EPI_ISL_606013|20 ···················· ···················· ···················· ···················· ···················· ····················
O 3082 hCoV-19/England/ALDP-A874CF/2020|EPI_ISL_606014|20 ···················· ···················· ···················· ···················· ···················· ····················
O 3083 hCoV-19/England/ALDP-A86E4A/2020|EPI_ISL_606017|20 ···················· ···················· ···················· ···················· ···················· ····················
O 3084 hCoV-19/England/ALDP-A871B9/2020|EPI_ISL_606018|20 ···················· ···················· ···················· ···················· ···················· ····················
O 3085 hCoV-19/England/ALDP-A875AE/2020|EPI_ISL_606019|20 ···················· ···················· ···················· ···················· ···················· ····················
O 3086 hCoV-19/England/ALDP-A873E0/2020|EPI_ISL_606020|20 ···················· ···················· ···················· ···················· ···················· ····················
O 3087 hCoV-19/England/ALDP-A865CD/2020|EPI_ISL_606021|20 ···················· ···················· ···················· ···················· ···················· ····················
O 3088 hCoV-19/England/ALDP-A87CB8/2020|EPI_ISL_606022|20 ···················· ···················· ···················· ···················· ···················· ····················
O 3089 hCoV-19/England/ALDP-A860F9/2020|EPI_ISL_606023|20 ···················· ···················· ···················· ···················· ···················· ····················
O 3090 hCoV-19/England/ALDP-A87C21/2020|EPI_ISL_606024|20 ···················· ···················· ···················· ···················· ···················· ····················
O 3091 hCoV-19/England/ALDP-A861C9/2020|EPI_ISL_606025|20 ···················· ···················· ···················· ···················· ···················· ····················
O 3092 hCoV-19/England/ALDP-A86F56/2020|EPI_ISL_606026|20 ···················· ···················· ···················· ···················· ···················· ····················
O 3093 hCoV-19/England/ALDP-A87B06/2020|EPI_ISL_606027|20 ···················· ···················· ···················· ···················· ···················· ····················
O 3094 hCoV-19/England/ALDP-A86D3E/2020|EPI_ISL_606028|20 ···················· ···················· ···················· ···················· ···················· ····················
O 3095 hCoV-19/England/ALDP-A8619C/2020|EPI_ISL_606029|20 ···················· ···················· ···················· ···················· ···················· ····················
O 3096 hCoV-19/England/ALDP-A86D6B/2020|EPI_ISL_606030|20 ···················· ···················· ···················· ···················· ···················· ····················
O 3097 hCoV-19/England/ALDP-A8716E/2020|EPI_ISL_606031|20 ···················· ···················· ···················· ···················· ···················· ····················
O 3098 hCoV-19/England/ALDP-A87A72/2020|EPI_ISL_606032|20 ···················· ·A·················· ···················· ···················· ···················· ····················
O 3099 hCoV-19/England/ALDP-A87D88/2020|EPI_ISL_606033|20 ···················· ···················· ···················· ···················· ···················· ····················
O 3100 hCoV-19/England/ALDP-A86457/2020|EPI_ISL_606034|20 ···················· ···················· ···················· ···················· ···················· ····················
O 3101 hCoV-19/England/ALDP-A8667F/2020|EPI_ISL_606035|20 ···················· ···················· ···················· ···················· ···················· ····················
O 3102 hCoV-19/England/ALDP-A87104/2020|EPI_ISL_606036|20 ···················· ···················· ···················· ···················· ···················· ····················
O 3103 hCoV-19/England/ALDP-A86E68/2020|EPI_ISL_606037|20 ···················· ···················· ···················· ···················· ···················· ····················
O 3104 hCoV-19/England/ALDP-A87720/2020|EPI_ISL_606038|20 ···················· ···················· ···················· ···················· ···················· ····················
O 3105 hCoV-19/England/ALDP-A86FED/2020|EPI_ISL_606039|20 ···················· ···················· ···················· ···················· ···················· ····················
O 3106 hCoV-19/England/ALDP-A86008/2020|EPI_ISL_606041|20 ···················· ···················· ···················· ···················· ···················· ····················
O 3107 hCoV-19/England/ALDP-A86150/2020|EPI_ISL_606042|20 ···················· ···················· ···················· ···················· ···················· ····················
O 3108 hCoV-19/England/ALDP-A87C9A/2020|EPI_ISL_606043|20 ···················· ···················· ···················· ···················· ···················· ····················
O 3109 hCoV-19/England/ALDP-A8722F/2020|EPI_ISL_606044|20 ···················· ···················· ···················· ···················· ···················· ····················
O 3110 hCoV-19/England/ALDP-A86CD7/2020|EPI_ISL_606045|20 ···················· ···················· ···················· ···················· ···················· ····················
O 3111 hCoV-19/England/ALDP-A864FD/2020|EPI_ISL_606047|20 ···················· ···················· ···················· ···················· ···················· ····················
O 3112 hCoV-19/England/ALDP-A87957/2020|EPI_ISL_606048|20 ···················· ···················· ···················· ···················· ···················· ····················
O 3113 hCoV-19/England/ALDP-A87368/2020|EPI_ISL_606049|20 ···················· ···················· ···················· ···················· ···················· ····················
O 3114 hCoV-19/England/ALDP-A86F0B/2020|EPI_ISL_606050|20 ···················· ···················· ···················· ···················· ···················· ····················
O 3115 hCoV-19/England/ALDP-A87D4C/2020|EPI_ISL_606051|20 ···················· ···················· ···················· ···················· ···················· ····················
O 3116 hCoV-19/England/ALDP-A87517/2020|EPI_ISL_606052|20 ···················· ·A·················· ···················· ···················· ···················· ··A·················
O 3117 hCoV-19/England/ALDP-A86527/2020|EPI_ISL_606053|20 ···················· ···················· ···················· ···················· ···················· ····················
O 3118 hCoV-19/England/ALDP-A8632D/2020|EPI_ISL_606054|20 ···················· ···················· ···················· ···················· ···················· ····················
O 3119 hCoV-19/England/ALDP-A870E9/2020|EPI_ISL_606056|20 ···················· ···················· ···················· ···················· ···················· ····················
O 3120 hCoV-19/England/ALDP-A87034/2020|EPI_ISL_606057|20 ···················· ···················· ···················· ···················· ···················· ····················
O 3121 hCoV-19/England/ALDP-A86C8C/2020|EPI_ISL_606058|20 ···················· ···················· ···················· ···················· ···················· ····················
O 3122 hCoV-19/England/ALDP-A87447/2020|EPI_ISL_606059|20 ···················· ···················· ···················· ···················· ···················· ····················
O 3123 hCoV-19/England/MILK-A7C76A/2020|EPI_ISL_607310|20 ···················· ···················· ···················· ···················· ···················· ····················
O 3124 hCoV-19/England/MILK-A7C7A6/2020|EPI_ISL_607311|20 ···················· ···················· ···················· ···················· ·····T·············· ····················
O 3125 hCoV-19/England/MILK-A7B507/2020|EPI_ISL_607313|20 ···················· ···················· ···················· ···················· ···················· ····················
O 3126 hCoV-19/England/MILK-A7BE1B/2020|EPI_ISL_607314|20 ···················· ···················· ···················· ···················· ·····T·············· ····················
O 3127 hCoV-19/England/MILK-A7C69A/2020|EPI_ISL_607315|20 ···················· ···················· ···················· ···················· ···················· ····················
O 3128 hCoV-19/England/MILK-A7B80E/2020|EPI_ISL_607317|20 ···················· ···················· ···················· ···················· ···················· ····················
O 3129 hCoV-19/England/MILK-A7B0D9/2020|EPI_ISL_607318|20 ···················· ···················· ···················· ···················· ···················· ····················
O 3130 hCoV-19/England/MILK-A7B886/2020|EPI_ISL_607319|20 ···················· ···················· ···················· ···················· ···················· ····················
O 3131 hCoV-19/England/MILK-A7B789/2020|EPI_ISL_607320|20 ···················· ···················· ···················· ···················· ···················· ····················
O 3132 hCoV-19/England/MILK-A7BD0F/2020|EPI_ISL_607321|20 ···················· ···················· ···················· ···················· ···················· ····················
O 3133 hCoV-19/England/MILK-A7BD69/2020|EPI_ISL_607324|20 ···················· ···················· ···················· ···················· ·····T·············· ····················
O 3134 hCoV-19/England/MILK-A7B015/2020|EPI_ISL_607325|20 ···················· ···················· ···················· ···················· ·····T·············· ····················

```
O 3135 hCoV-19/England/MILK-A7BF63/2020|EPI_ISL_607328|20 ···················· ···················· ····················· ···················· ···················· ····················
O 3136 hCoV-19/England/MILK-A7C533/2020|EPI_ISL_607330|20 ···················· ···················· ····················· ···················· ···················· ····················
O 3137 hCoV-19/England/MILK-A7B1D6/2020|EPI_ISL_607331|20 ···················· ···················· ····················· ···················· ···················· ····················
O 3138 hCoV-19/England/MILK-A7BF90/2020|EPI_ISL_607332|20 ···················· ···················· ····················· ···················· ······T············· ····················
O 3139 hCoV-19/England/MILK-A7C83A/2020|EPI_ISL_607333|20 ···················· ···················· ····················· ···················· ···················· ····················
O 3140 hCoV-19/England/MILK-A7CC4D/2020|EPI_ISL_607335|20 ···················· ···················· ····················· ···················· ···················· ····················
O 3141 hCoV-19/England/MILK-A7AFFB/2020|EPI_ISL_607336|20 ···················· ···················· ····················· ···················· ···················· ····················
O 3142 hCoV-19/England/MILK-A7BEA2/2020|EPI_ISL_607337|20 ···················· ···················· ····················· ···················· ···················· ····················
O 3143 hCoV-19/England/MILK-A7CBC8/2020|EPI_ISL_607339|20 ···················· ···················· ····················· ···················· ···················· ····················
O 3144 hCoV-19/England/MILK-A7CAE9/2020|EPI_ISL_607344|20 ···················· ···················· ····················· ···················· ···················· ····················
O 3145 hCoV-19/England/MILK-A7BA80/2020|EPI_ISL_607346|20 ···················· ·A·················· ····················· ···················· ···················· ···A················
O 3146 hCoV-19/England/MILK-A7B473/2020|EPI_ISL_607347|20 ···················· ···················· ····················· ···················· ···················· ····················
O 3147 hCoV-19/England/MILK-A7C14E/2020|EPI_ISL_607348|20 ···················· ···················· ····················· ···················· ···················· ····················
O 3148 hCoV-19/England/MILK-A7BA17/2020|EPI_ISL_607349|20 ···················· ···················· ····················· ···················· ···················· ····················
O 3149 hCoV-19/England/MILK-A7B9FC/2020|EPI_ISL_607350|20 ···················· ·A·················· ····················· ···················· ···················· ···A················
O 3150 hCoV-19/England/MILK-A7CCA7/2020|EPI_ISL_607351|20 ···················· ···················· ····················· ···················· ······T············· ····················
O 3151 hCoV-19/England/MILK-A7B710/2020|EPI_ISL_607352|20 ···················· ·A·················· ····················· ···················· ···················· ···A················
O 3152 hCoV-19/England/MILK-A7BEDF/2020|EPI_ISL_607354|20 ···················· ···················· ····················· ···················· ···················· ····················
O 3153 hCoV-19/England/MILK-A7B112/2020|EPI_ISL_607355|20 ···················· ···················· ····················· ···················· ···················· ····················
O 3154 hCoV-19/England/MILK-A7BA71/2020|EPI_ISL_607356|20 ···················· ···················· ····················· ···················· ···················· ····················
O 3155 hCoV-19/England/MILK-A7BB32/2020|EPI_ISL_607358|20 ···················· ·A·················· ····················· ···················· ···················· ···A················
O 3156 hCoV-19/England/MILK-A7B938/2020|EPI_ISL_607359|20 ···················· ···················· ····················· ···················· ···················· ····················
O 3157 hCoV-19/England/MILK-A7B8A4/2020|EPI_ISL_607360|20 ···················· ···················· ····················· ···················· ···················· ····················
O 3158 hCoV-19/England/MILK-A7CC5C/2020|EPI_ISL_607361|20 ···················· ···················· ····················· ···················· ···················· ····················
O 3159 hCoV-19/England/MILK-A7B21F/2020|EPI_ISL_607362|20 ···················· ···················· ····················· ···················· ···················· ····················
O 3160 hCoV-19/England/MILK-A7B18B/2020|EPI_ISL_607363|20 ···················· ···················· ····················· ···················· ···················· ····················
O 3161 hCoV-19/England/MILK-A7BCB7/2020|EPI_ISL_607364|20 ···················· ···················· ····················· ···················· ···················· ····················
O 3162 hCoV-19/England/MILK-A7B3EF/2020|EPI_ISL_607365|20 ···················· ···················· ····················· ···················· ···················· ····················
O 3163 hCoV-19/England/MILK-A7C18A/2020|EPI_ISL_607366|20 ···················· ···················· ····················· ···················· ···················· ····················
O 3164 hCoV-19/England/MILK-A7BD96/2020|EPI_ISL_607367|20 ···················· ···················· ····················· ···················· ···················· ····················
O 3166 hCoV-19/England/MILK-A7BEFD/2020|EPI_ISL_607369|20 ···················· ···················· ····················· ···················· ···················· ····················
O 3167 hCoV-19/England/MILK-A7B9DE/2020|EPI_ISL_607370|20 ···················· ···················· ····················· ···················· ···················· ····················
O 3168 hCoV-19/England/MILK-A7B464/2020|EPI_ISL_607371|20 ···················· ·A·················· ····················· ···················· ···················· ···A················
O 3169 hCoV-19/England/MILK-A7BFFA/2020|EPI_ISL_607372|20 ···················· ···················· ····················· ···················· ···················· ····················
O 3170 hCoV-19/England/MILK-A7BCF3/2020|EPI_ISL_607374|20 ···················· ···················· ····················· ···················· ···················· ····················
O 3171 hCoV-19/England/MILK-A7B8D1/2020|EPI_ISL_607376|20 ···················· ···················· ····················· ···················· ···················· ····················
O 3172 hCoV-19/England/MILK-A7B82C/2020|EPI_ISL_607377|20 ···················· ···················· ····················· ···················· ···················· ····················
O 3173 hCoV-19/England/MILK-A7C867/2020|EPI_ISL_607378|20 ···················· ···················· ····················· ···················· ···················· ····················
O 3174 hCoV-19/England/MILK-A7AFEC/2020|EPI_ISL_607379|20 ···················· ···················· ····················· ···················· ······T············· ····················
O 3175 hCoV-19/England/MILK-A7CC7A/2020|EPI_ISL_607380|20 ···················· ···················· ····················· ···················· ···················· ····················
O 3176 hCoV-19/England/MILK-A7C955/2020|EPI_ISL_607381|20 ···················· ···················· ····················· ···················· ···················· ····················
O 3177 hCoV-19/England/MILK-A7CC98/2020|EPI_ISL_607382|20 ···················· ···················· ····················· ···················· ···················· ····················
O 3178 hCoV-19/England/MILK-A7CB22/2020|EPI_ISL_607383|20 ···················· ·A·················· ····················· ···················· ···················· ···A················
O 3179 hCoV-19/England/MILK-A7C779/2020|EPI_ISL_607384|20 ···················· ···················· ····················· ···················· ···················· ····················
O 3180 hCoV-19/England/MILK-A7B90B/2020|EPI_ISL_607385|20 ···················· ···················· ····················· ···················· ···················· ····················
O 3181 hCoV-19/England/MILK-A7C9DD/2020|EPI_ISL_607386|20 ···················· ···················· ····················· ···················· ···················· ····················
O 3182 hCoV-19/England/MILK-A7B992/2020|EPI_ISL_607387|20 ···················· ···················· ····················· ···················· ···················· ····················
O 3183 hCoV-19/England/MILK-A7BAEA/2020|EPI_ISL_607388|20 ···················· ···················· ····················· ···················· ···················· ····················
O 3184 hCoV-19/England/MILK-A7CC10/2020|EPI_ISL_607389|20 ···················· ···················· ····················· ···················· ····T··············· ····················
O 3185 hCoV-19/England/ALDP-A68B47/2020|EPI_ISL_608640|20 ···················· ···················· ····················· ···················· ···················· ····················
O 3186 hCoV-19/England/CAMC-A64B2D/2020|EPI_ISL_608641|20 ···················· ···················· ····················· ···················· ····T··············· ····················
O 3187 hCoV-19/England/CAMC-A65940/2020|EPI_ISL_608642|20 ···················· ···················· ····················· ···················· ···················· ····················
O 3188 hCoV-19/England/CAMC-A65078/2020|EPI_ISL_608643|20 ···················· ···················· ····················· ···················· ···················· ····················
O 3189 hCoV-19/England/CAMC-A64F6D/2020|EPI_ISL_608644|20 ···················· ···················· ····················· ···················· ···················· ····················
O 3190 hCoV-19/England/CAMC-A64B69/2020|EPI_ISL_608645|20 ···················· ···················· ····················· ···················· ···················· ····················
O 3191 hCoV-19/England/ALDP-A68EE4/2020|EPI_ISL_608646|20 ···················· ···················· ····················· ···················· ······T············· ····················
O 3192 hCoV-19/England/ALDP-A687AD/2020|EPI_ISL_608647|20 ···················· ···················· ····················· ···················· ···················· ····················
O 3193 hCoV-19/England/ALDP-A687E9/2020|EPI_ISL_608648|20 ···················· ···················· ····················· ···················· ···················· ····················
O 3194 hCoV-19/England/ALDP-A691DB/2020|EPI_ISL_608649|20 ···················· ·A·······A·········· ····················· ···················· ···················· ····················
O 3195 hCoV-19/England/CAMC-A6551F/2020|EPI_ISL_608650|20 ···················· ···················· ····················· ···················· ···················· ····················
O 3196 hCoV-19/England/ALDP-A67B93/2020|EPI_ISL_608652|20 ···················· ···················· ····················· ···················· ···················· ····················
O 3197 hCoV-19/England/CAMC-A65A6B/2020|EPI_ISL_608653|20 ···················· ···················· ····················· ···················· ···················· ····················
O 3198 hCoV-19/England/CAMC-A65597/2020|EPI_ISL_608654|20 ···················· ···················· ····················· ···················· ···················· ····················
O 3199 hCoV-19/England/ALDP-A68C62/2020|EPI_ISL_608655|20 ···················· ···················· ····················· ···················· ···················· ····················
O 3200 hCoV-19/England/CAMC-A6596E/2020|EPI_ISL_608656|20 ···················· ···················· ····················· ···················· ···················· ····················
O 3201 hCoV-19/England/ALDP-A68EB7/2020|EPI_ISL_608657|20 ···················· ···················· ····················· ···················· ···················· ····················
O 3202 hCoV-19/England/ALDP-A69126/2020|EPI_ISL_608658|20 ···················· ···················· ····················· ···················· ···················· ····················
O 3203 hCoV-19/England/ALDP-A68E20/2020|EPI_ISL_608659|20 ···················· ·A·················· ····················· ···················· ···················· ····················
O 3204 hCoV-19/England/ALDP-A683E5/2020|EPI_ISL_608661|20 ···················· ···················· ····················· ···················· ···················· ····················
O 3205 hCoV-19/England/ALDP-A688B9/2020|EPI_ISL_608663|20 ···················· ·A·················· ····················· ···················· ···················· ····················
O 3206 hCoV-19/England/CAMC-A65184/2020|EPI_ISL_608664|20 ···················· ···················· ····················· ···················· ···················· ····················
O 3207 hCoV-19/England/ALDP-A6837C/2020|EPI_ISL_608665|20 ···················· ···················· ····················· ···················· ···················· ····················
O 3208 hCoV-19/England/CAMC-A64ED9/2020|EPI_ISL_608666|20 ···················· ···················· ····················· ···················· ···················· ····················
O 3209 hCoV-19/England/ALDP-A68C08/2020|EPI_ISL_608667|20 ···················· ···················· ····················· ···················· ···················· ····················
O 3210 hCoV-19/England/ALDP-A68E02/2020|EPI_ISL_608668|20 ···················· ···················· ····················· ···················· ···················· ····················
O 3211 hCoV-19/England/ALDP-A68B74/2020|EPI_ISL_608669|20 ···················· ···················· ····················· ···················· ···················· ····················
O 3212 hCoV-19/England/ALDP-A68CAE/2020|EPI_ISL_608670|20 ···················· ···················· ····················· ···················· ···················· ····················
O 3213 hCoV-19/England/ALDP-A688E6/2020|EPI_ISL_608675|20 ···················· ···················· ····················· ···················· ···················· ····················
O 3214 hCoV-19/England/ALDP-A68B65/2020|EPI_ISL_608676|20 ···················· ···················· ····················· ···················· ···················· ····················
O 3215 hCoV-19/England/ALDP-A67EF4/2020|EPI_ISL_608677|20 ···················· ···················· ····················· ···················· ···················· ····················
```

| | | | | | | | | |
|---|---|---|---|---|---|---|---|---|
| O | 3216 | hCoV-19/England/CAMC-A64C2A/2020\|EPI_ISL_608678\|20 | ···················· | ···················· | ···················· | ···················· | ···················· | ···················· |
| O | 3217 | hCoV-19/England/CAMC-A658DA/2020\|EPI_ISL_608679\|20 | ···················· | ·A·················· | ···················· | ···················· | ···················· | ···················· |
| O | 3218 | hCoV-19/England/ALDP-A68F87/2020\|EPI_ISL_608680\|20 | ···················· | ···················· | ···················· | ···················· | ···················· | ···················· |
| O | 3219 | hCoV-19/England/CAMC-A658AD/2020\|EPI_ISL_608683\|20 | ···················· | ···················· | ···················· | ···················· | ···················· | ···················· |
| O | 3220 | hCoV-19/England/ALDP-A68567/2020\|EPI_ISL_608684\|20 | ···················· | ···················· | ···················· | ···················· | ···················· | ···················· |
| O | 3221 | hCoV-19/England/ALDP-A6886E/2020\|EPI_ISL_608685\|20 | ···················· | ···················· | ···················· | ···················· | ···················· | ···················· |
| O | 3222 | hCoV-19/England/ALDP-A68C53/2020\|EPI_ISL_608686\|20 | ···················· | ···················· | ···················· | ···················· | ···················· | ···················· |
| O | 3223 | hCoV-19/England/ALDP-A67B84/2020\|EPI_ISL_608687\|20 | ···················· | ···················· | ···················· | ···················· | ···················· | ···················· |
| O | 3224 | hCoV-19/England/CAMC-A64F03/2020\|EPI_ISL_608688\|20 | ···················· | ···················· | ···················· | ···················· | ···················· | ···················· |
| O | 3225 | hCoV-19/England/CAMC-A65412/2020\|EPI_ISL_608689\|20 | ···················· | ···················· | ···················· | ···················· | ···················· | ···················· |
| O | 3226 | hCoV-19/Wales/ALDP-A6836D/2020\|EPI_ISL_608690\|2020 | ···················· | ···················· | ···················· | ···················· | ···················· | ···················· |
| O | 3227 | hCoV-19/England/ALDP-A68770/2020\|EPI_ISL_608691\|20 | ···················· | ···················· | ···················· | ···················· | ···················· | ···················· |
| O | 3228 | hCoV-19/England/ALDP-A68C80/2020\|EPI_ISL_608692\|20 | ···················· | ···················· | ···················· | ···················· | ···················· | ···················· |
| O | 3229 | hCoV-19/England/CAMC-A64BA5/2020\|EPI_ISL_608693\|20 | ···················· | ···················· | ···················· | ···················· | ···················· | ···················· |
| O | 3230 | hCoV-19/England/ALDP-A6892F/2020\|EPI_ISL_608694\|20 | ···················· | ···················· | ···················· | ···················· | ···················· | ···················· |
| O | 3231 | hCoV-19/England/CAMC-A64F21/2020\|EPI_ISL_608695\|20 | ···················· | ···················· | ···················· | ···················· | ···················· | ···················· |
| O | 3232 | hCoV-19/England/CAMC-A65685/2020\|EPI_ISL_608696\|20 | ···················· | ···················· | ···················· | ···················· | ···················· | ···················· |
| O | 3233 | hCoV-19/England/MILK-A6B230/2020\|EPI_ISL_608697\|20 | ···················· | ···················· | ···················· | ···················· | ···················· | ···················· |
| O | 3234 | hCoV-19/England/MILK-A6B740/2020\|EPI_ISL_608698\|20 | ···················· | ···················· | ···················· | ···················· | ···················· | ···················· |
| O | 3235 | hCoV-19/England/MILK-A6B652/2020\|EPI_ISL_608699\|20 | ···················· | ···················· | ···················· | ···················· | ···················· | ···················· |
| O | 3236 | hCoV-19/England/MILK-A6B31F/2020\|EPI_ISL_608700\|20 | ···················· | ···················· | ···················· | ···················· | ···················· | ···················· |
| O | 3237 | hCoV-19/England/MILK-A6B1F7/2020\|EPI_ISL_608701\|20 | ···················· | ···················· | ···················· | ···················· | ···················· | ···················· |
| O | 3238 | hCoV-19/England/MILK-A6B133/2020\|EPI_ISL_608702\|20 | ···················· | ···················· | ···················· | ···················· | ···················· | ···················· |
| O | 3239 | hCoV-19/England/MILK-A6B76E/2020\|EPI_ISL_608703\|20 | ···················· | ···················· | ···················· | ···················· | ···················· | ···················· |
| O | 3240 | hCoV-19/England/MILK-A6AF1C/2020\|EPI_ISL_608705\|20 | ···················· | ···················· | ···················· | ···················· | ···················· | ···················· |
| O | 3241 | hCoV-19/England/MILK-A6BBCC/2020\|EPI_ISL_608706\|20 | ···················· | ···················· | ···················· | ···················· | ···················· | ···················· |
| O | 3242 | hCoV-19/England/MILK-A6B0DC/2020\|EPI_ISL_608707\|20 | ···················· | ·A·················· | ···················· | ···················· | ···················· | ··A················· |
| O | 3243 | hCoV-19/England/MILK-A6B1CA/2020\|EPI_ISL_608708\|20 | ···················· | ···················· | ···················· | ···················· | ···················· | ···················· |
| O | 3244 | hCoV-19/England/MILK-A69CE9/2020\|EPI_ISL_608709\|20 | ···················· | ···················· | ···················· | ···················· | ···················· | ···················· |
| O | 3245 | hCoV-19/England/MILK-A6B388/2020\|EPI_ISL_608710\|20 | ···················· | ···················· | ···················· | ···················· | ···················· | ···················· |
| O | 3246 | hCoV-19/England/MILK-A6B4EF/2020\|EPI_ISL_608711\|20 | ···················· | ···················· | ···················· | ···················· | ···················· | ···················· |
| O | 3247 | hCoV-19/England/MILK-A6AFB2/2020\|EPI_ISL_608713\|20 | ···················· | ···················· | ···················· | ···················· | ···················· | ···················· |
| O | 3248 | hCoV-19/England/MILK-A69D7D/2020\|EPI_ISL_608714\|20 | ···················· | ···················· | ···················· | ···················· | ···················· | ···················· |
| O | 3249 | hCoV-19/England/MILK-A6BB08/2020\|EPI_ISL_608716\|20 | ···················· | ···················· | ···················· | ···················· | ···················· | ···················· |
| O | 3250 | hCoV-19/England/MILK-A6AF0D/2020\|EPI_ISL_608717\|20 | ···················· | ···················· | ···················· | ···················· | ···················· | ···················· |
| O | 3251 | hCoV-19/England/MILK-A6BA38/2020\|EPI_ISL_608718\|20 | ···················· | ···················· | ···················· | ···················· | ······T············· | ···················· |
| O | 3252 | hCoV-19/England/MILK-A6ABEB/2020\|EPI_ISL_608719\|20 | ···················· | ···················· | ···················· | ···················· | ···················· | ···················· |
| O | 3253 | hCoV-19/USA/VA-DCLS-1681/2020\|EPI_ISL_609971\|2020- | ···················· | ···················· | ···················· | ···················· | ···················· | ···················· |
| O | 3254 | hCoV-19/USA/VA-DCLS-1682/2020\|EPI_ISL_609972\|2020- | ···················· | ···················· | ···················· | ···················· | ···················· | ···················· |
| O | 3255 | hCoV-19/USA/VA-DCLS-1683/2020\|EPI_ISL_609973\|2020- | ···················· | ·A·················· | ···················· | ···················· | ···················· | ···················· |
| O | 3256 | hCoV-19/USA/VA-DCLS-1684/2020\|EPI_ISL_609974\|2020- | ···················· | ···················· | ···················· | ···················· | ···················· | ···················· |
| O | 3257 | hCoV-19/USA/VA-DCLS-1685/2020\|EPI_ISL_609975\|2020- | ···················· | ···················· | ···················· | ···················· | ···················· | ···················· |
| O | 3258 | hCoV-19/USA/VA-DCLS-1686/2020\|EPI_ISL_609976\|2020- | ···················· | ···················· | ···················· | ···················· | ···················· | ···················· |
| O | 3259 | hCoV-19/USA/VA-DCLS-1691/2020\|EPI_ISL_609977\|2020- | ···················· | ···················· | ···················· | ···················· | ···················· | ···················· |
| O | 3260 | hCoV-19/USA/VA-DCLS-1696/2020\|EPI_ISL_609978\|2020- | ···················· | ···················· | ···················· | ···················· | ···················· | ···················· |
| O | 3261 | hCoV-19/USA/VA-DCLS-1698/2020\|EPI_ISL_609979\|2020- | ···················· | ···················· | ···················· | ···················· | ···················· | ···················· |
| O | 3262 | hCoV-19/USA/VA-DCLS-1699/2020\|EPI_ISL_609980\|2020- | ···················· | ···················· | ···················· | ···················· | ···················· | ···················· |
| O | 3263 | hCoV-19/USA/VA-DCLS-1701/2020\|EPI_ISL_609981\|2020- | ···················· | ···················· | ···················· | ···················· | ···················· | ···················· |
| O | 3264 | hCoV-19/USA/VA-DCLS-1702/2020\|EPI_ISL_609982\|2020- | ···················· | ···················· | ···················· | ···················· | ···················· | ···················· |
| O | 3265 | hCoV-19/USA/VA-DCLS-1704/2020\|EPI_ISL_609983\|2020- | ···················· | ···················· | ···················· | ···················· | ···················· | ···················· |
| O | 3266 | hCoV-19/USA/VA-DCLS-1812/2020\|EPI_ISL_609984\|2020- | ···················· | ·A·················· | ···················· | ···················· | ···················· | ···················· |
| O | 3267 | hCoV-19/USA/VA-DCLS-1814/2020\|EPI_ISL_609985\|2020- | ···················· | ·A·················· | ···················· | ···················· | ···················· | ···················· |
| O | 3268 | hCoV-19/USA/VA-DCLS-1830/2020\|EPI_ISL_609986\|2020- | ···················· | ·A·················· | ···················· | ···················· | ···················· | ···················· |
| O | 3269 | hCoV-19/Italy/LAZ-INMI-81/2020\|EPI_ISL_609989\|2020 | ···················· | ···················· | ···················· | ···················· | ···················· | ···················· |
| O | 3270 | hCoV-19/Italy/LAZ-INMI-82/2020\|EPI_ISL_609990\|2020 | ···················· | ···················· | ···················· | ···················· | ···················· | ···················· |
| O | 3271 | hCoV-19/Italy/LAZ-INMI-83/2020\|EPI_ISL_609991\|2020 | ···················· | ·A·················· | ···················· | ···················· | ···················· | ···················· |
| O | 3272 | hCoV-19/Italy/LAZ-INMI-84/2020\|EPI_ISL_609992\|2020 | ···················· | ···················· | ···················· | ···················· | ······T············· | ···················· |
| O | 3273 | hCoV-19/Italy/LAZ-INMI-85/2020\|EPI_ISL_609993\|2020 | ···················· | ···················· | ···················· | ···················· | ······T············· | ···················· |
| O | 3274 | hCoV-19/Italy/LAZ-INMI-86/2020\|EPI_ISL_609994\|2020 | ···················· | ···················· | ···················· | ···················· | ···················· | ···················· |
| O | 3275 | hCoV-19/Italy/LAZ-INMI-87/2020\|EPI_ISL_609995\|2020 | ···················· | ···················· | ···················· | ···················· | ······T············· | ···················· |
| O | 3278 | hCoV-19/Italy/LAZ-INMI-90/2020\|EPI_ISL_609998\|2020 | ···················· | ···················· | ···················· | ···················· | ······T············· | ···················· |
| O | 3279 | hCoV-19/Italy/LAZ-INMI-91/2020\|EPI_ISL_609999\|2020 | ···················· | ···················· | ···················· | ···················· | ······T············· | ···················· |
| O | 3280 | hCoV-19/England/ALDP-A92379/2020\|EPI_ISL_610953\|20 | ···················· | ···················· | ···················· | ···················· | ···················· | ···················· |
| O | 3281 | hCoV-19/England/ALDP-A9393A/2020\|EPI_ISL_610954\|20 | ···················· | ···················· | ···················· | ···················· | ···················· | ···················· |
| O | 3282 | hCoV-19/England/ALDP-A92449/2020\|EPI_ISL_610955\|20 | ···················· | ···················· | ···················· | ···················· | ···················· | ···················· |
| O | 3283 | hCoV-19/England/ALDP-A923F1/2020\|EPI_ISL_610957\|20 | ···················· | ···················· | ···················· | ···················· | ···················· | ···················· |
| O | 3284 | hCoV-19/England/ALDP-A93FCF/2020\|EPI_ISL_610958\|20 | ···················· | ·A·················· | ···················· | ···················· | ···················· | ···················· |
| O | 3285 | hCoV-19/England/ALDP-A928F2/2020\|EPI_ISL_610960\|20 | ···················· | ···················· | ···················· | ···················· | ···················· | ···················· |
| O | 3286 | hCoV-19/England/ALDP-A946AB/2020\|EPI_ISL_610961\|20 | ···················· | ···················· | ···················· | ···················· | ···················· | ···················· |
| O | 3287 | hCoV-19/England/ALDP-A923D3/2020\|EPI_ISL_610962\|20 | ···················· | ···················· | ···················· | ···················· | ···················· | ···················· |
| O | 3288 | hCoV-19/England/ALDP-A93994/2020\|EPI_ISL_610963\|20 | ···················· | ···················· | ···················· | ···················· | ···················· | ···················· |
| O | 3289 | hCoV-19/England/ALDP-A93B43/2020\|EPI_ISL_610964\|20 | ···················· | ···················· | ···················· | ···················· | ···················· | ···················· |
| O | 3290 | hCoV-19/England/ALDP-A93B9E/2020\|EPI_ISL_610965\|20 | ···················· | ···················· | ···················· | ···················· | ···················· | ···················· |
| O | 3291 | hCoV-19/England/ALDP-A96E65/2020\|EPI_ISL_610966\|20 | ···················· | ···················· | ···················· | ···················· | ···················· | ···················· |
| O | 3292 | hCoV-19/England/ALDP-A92DF3/2020\|EPI_ISL_610967\|20 | ···················· | ···················· | ···················· | ···················· | ···················· | ···················· |
| O | 3293 | hCoV-19/England/ALDP-A947E4/2020\|EPI_ISL_610968\|20 | ···················· | ···················· | ···················· | ···················· | ···················· | ··A················· |
| O | 3294 | hCoV-19/England/ALDP-A92713/2020\|EPI_ISL_610972\|20 | ···················· | ···················· | ···················· | ···················· | ···················· | ···················· |
| O | 3295 | hCoV-19/England/ALDP-A92AA1/2020\|EPI_ISL_610973\|20 | ···················· | ···················· | ···················· | ···················· | ···················· | ···················· |
| O | 3296 | hCoV-19/England/ALDP-A9409E/2020\|EPI_ISL_610974\|20 | ············T······· | ···················· | ···················· | ···················· | ···················· | ···················· |
| O | 3297 | hCoV-19/England/ALDP-A93F83/2020\|EPI_ISL_610975\|20 | ···················· | ···················· | ···················· | ···················· | ···················· | ···················· |

| | | | | | | | |
|---|---|---|---|---|---|---|---|
| O | 3298 hCoV-19/England/ALDP-A9234C/2020\|EPI_ISL_610976\|20 | .................... | .................... | ..................... | .................... | .................... | .................... |
| O | 3299 hCoV-19/England/ALDP-A926AD/2020\|EPI_ISL_610977\|20 | .................... | .................... | ..................... | .................... | .................... | .................... |
| O | 3300 hCoV-19/England/ALDP-A92977/2020\|EPI_ISL_610978\|20 | .................... | .................... | ..................... | .................... | .................... | .................... |
| O | 3301 hCoV-19/England/ALDP-A92397/2020\|EPI_ISL_610979\|20 | .................... | .................... | ..................... | .................... | .................... | .................... |
| O | 3302 hCoV-19/England/ALDP-A93EC2/2020\|EPI_ISL_610981\|20 | .................... | .................... | ..................... | .................... | .................... | .................... |
| O | 3303 hCoV-19/England/ALDP-A936AC/2020\|EPI_ISL_610982\|20 | .................... | .................... | ..................... | .................... | .................... | .................... |
| O | 3304 hCoV-19/England/ALDP-A9278C/2020\|EPI_ISL_610983\|20 | .................... | .................... | ..................... | .................... | .................... | .................... |
| O | 3305 hCoV-19/England/ALDP-A93BAD/2020\|EPI_ISL_610985\|20 | .................... | .................... | ..................... | .................... | .................... | .................... |
| O | 3306 hCoV-19/England/ALDP-A92D02/2020\|EPI_ISL_610986\|20 | .................... | .................... | ..................... | .................... | .................... | .................... |
| O | 3307 hCoV-19/England/ALDP-A9294A/2020\|EPI_ISL_610987\|20 | .................... | .................... | ..................... | .................... | .................... | .................... |
| O | 3308 hCoV-19/England/ALDP-A92A38/2020\|EPI_ISL_610988\|20 | .................... | .................... | ..................... | .................... | .................... | .................... |
| O | 3309 hCoV-19/England/ALDP-A92E5A/2020\|EPI_ISL_610990\|20 | .................... | .................... | ..................... | .................... | .................... | .................... |
| O | 3310 hCoV-19/England/ALDP-A93E2C/2020\|EPI_ISL_610991\|20 | .................... | .A.................. | ..................... | .................... | .................... | ..A................. |
| O | 3311 hCoV-19/England/ALDP-A92BEA/2020\|EPI_ISL_610992\|20 | .................... | .................... | ..................... | .................... | .................... | .................... |
| O | 3312 hCoV-19/England/ALDP-A93E59/2020\|EPI_ISL_610993\|20 | .................... | .A.................. | ..................... | .................... | .................... | ..A................. |
| O | 3313 hCoV-19/England/ALDP-A92607/2020\|EPI_ISL_610994\|20 | .................... | .................... | ..................... | .................... | .................... | .................... |
| O | 3314 hCoV-19/England/ALDP-A92C8D/2020\|EPI_ISL_610995\|20 | .................... | .................... | ..................... | .................... | .................... | .................... |
| O | 3315 hCoV-19/England/ALDP-A9483C/2020\|EPI_ISL_610996\|20 | .................... | .................... | ..................... | .................... | .................... | .................... |
| O | 3316 hCoV-19/England/ALDP-A92968/2020\|EPI_ISL_610997\|20 | .................... | .................... | ..................... | .................... | .................... | .................... |
| O | 3317 hCoV-19/England/PORT-2D21C6/2020\|EPI_ISL_613284\|20 | .................... | .................... | ..................... | .................... | .................... | .................... |
| O | 3318 hCoV-19/England/PORT-2D220F/2020\|EPI_ISL_613285\|20 | .................... | .................... | ..................... | .................... | .................... | .................... |
| O | 3319 hCoV-19/England/PORT-2D46C5/2020\|EPI_ISL_613290\|20 | .................... | .................... | ..................... | .................... | ......T............. | .................... |
| O | 3320 hCoV-19/England/PORT-2D46D4/2020\|EPI_ISL_613291\|20 | .................... | .................... | ..................... | .................... | ......T............. | .................... |
| O | 3321 hCoV-19/England/PORT-2D473B/2020\|EPI_ISL_613292\|20 | .................... | .................... | ..................... | .................... | ......T............. | .................... |
| O | 3322 hCoV-19/England/PORT-2D4759/2020\|EPI_ISL_613293\|20 | .................... | .................... | ..................... | .................... | ......T............. | .................... |
| O | 3323 hCoV-19/England/PORT-2D47C2/2020\|EPI_ISL_613294\|20 | .................... | .................... | ..................... | .................... | ......T............. | .................... |
| O | 3324 hCoV-19/England/PORT-2D47D1/2020\|EPI_ISL_613295\|20 | .................... | .................... | ..................... | .................... | ......T............. | .................... |
| O | 3325 hCoV-19/England/PORT-2D4892/2020\|EPI_ISL_613296\|20 | .................... | .................... | ..................... | .................... | ......T............. | .................... |
| O | 3326 hCoV-19/England/PORT-2D4908/2020\|EPI_ISL_613297\|20 | .................... | .................... | ..................... | .................... | ......T............. | .................... |
| O | 3327 hCoV-19/England/PORT-2D49EA/2020\|EPI_ISL_613298\|20 | .................... | .................... | ..................... | .................... | .................... | .................... |
| O | 3328 hCoV-19/England/PORT-2F5752/2020\|EPI_ISL_613299\|20 | .................... | .................... | ..................... | .................... | .................... | .................... |
| O | 3329 hCoV-19/England/ALDP-A8766F/2020\|EPI_ISL_606010\|20 | .................... | .................... | ..................... | .................... | .................... | .................... |
| O | 3330 hCoV-19/England/ALDP-A876F6/2020\|EPI_ISL_606011\|20 | .................... | .................... | ..................... | .................... | .................... | .................... |
| O | 3331 hCoV-19/England/ALDP-A87D2E/2020\|EPI_ISL_606015\|20 | .................... | .................... | ..................... | .................... | .................... | .................... |
| O | 3332 hCoV-19/England/ALDP-A87B8E/2020\|EPI_ISL_606060\|20 | .................... | .................... | ..................... | .................... | .................... | .................... |
| O | 3333 hCoV-19/England/ALDP-A861F6/2020\|EPI_ISL_606061\|20 | .................... | .................... | ..................... | .................... | .................... | .................... |
| O | 3334 hCoV-19/England/ALDP-A87D5B/2020\|EPI_ISL_606062\|20 | .................... | .................... | ..................... | .................... | .................... | .................... |
| O | 3335 hCoV-19/England/ALDP-A8773F/2020\|EPI_ISL_606063\|20 | .................... | .................... | ..................... | .................... | .................... | .................... |
| O | 3336 hCoV-19/England/ALDP-A86D5C/2020\|EPI_ISL_606064\|20 | .................... | .................... | ..................... | .................... | .................... | ..A................. |
| O | 3337 hCoV-19/England/ALDP-A864EE/2020\|EPI_ISL_606065\|20 | .................... | .................... | ..................... | .................... | .................... | .................... |
| O | 3338 hCoV-19/England/ALDP-A86E86/2020\|EPI_ISL_606066\|20 | .................... | .................... | ..................... | .................... | .................... | .................... |
| O | 3339 hCoV-19/England/ALDP-A874DE/2020\|EPI_ISL_606067\|20 | .................... | .................... | ..................... | .................... | .................... | .................... |
| O | 3340 hCoV-19/England/ALDP-A879B1/2020\|EPI_ISL_606068\|20 | .................... | .................... | ..................... | .................... | .................... | .................... |
| O | 3341 hCoV-19/England/ALDP-A86448/2020\|EPI_ISL_606069\|20 | .................... | .................... | ..................... | .................... | .................... | .................... |
| O | 3342 hCoV-19/England/ALDP-A86F65/2020\|EPI_ISL_606070\|20 | .................... | .................... | ..................... | .................... | .................... | .................... |
| O | 3343 hCoV-19/England/ALDP-A863A5/2020\|EPI_ISL_606071\|20 | .................... | .................... | ..................... | .................... | .................... | .................... |
| O | 3344 hCoV-19/England/ALDP-A8623F/2020\|EPI_ISL_606072\|20 | .................... | .................... | ..................... | .................... | .................... | .................... |
| O | 3345 hCoV-19/England/ALDP-A87C5E/2020\|EPI_ISL_606074\|20 | .................... | .................... | ..................... | .................... | .................... | .................... |
| O | 3346 hCoV-19/England/ALDP-A86C7D/2020\|EPI_ISL_606075\|20 | .................... | .................... | ..................... | .................... | .................... | .................... |
| O | 3347 hCoV-19/England/ALDP-A861D8/2020\|EPI_ISL_606076\|20 | .................... | .................... | ..................... | .................... | .................... | .................... |
| O | 3348 hCoV-19/England/ALDP-A863B4/2020\|EPI_ISL_606077\|20 | .................... | .................... | ..................... | .................... | .................... | .................... |
| O | 3349 hCoV-19/England/ALDP-A86EFF/2020\|EPI_ISL_606079\|20 | .................... | .................... | ..................... | .................... | .................... | .................... |
| O | 3350 hCoV-19/England/ALDP-A87D00/2020\|EPI_ISL_606080\|20 | .................... | .................... | ..................... | .................... | .................... | .................... |
| O | 3351 hCoV-19/England/ALDP-A87D79/2020\|EPI_ISL_606081\|20 | .................... | .................... | ..................... | .................... | ......T............. | .................... |
| O | 3352 hCoV-19/England/ALDP-A86D7A/2020\|EPI_ISL_606082\|20 | .................... | .A.................. | ..................... | .................... | .................... | ..A................. |
| O | 3353 hCoV-19/England/ALDP-A86DC5/2020\|EPI_ISL_606083\|20 | .................... | .................... | ..................... | .................... | .................... | .................... |
| O | 3354 hCoV-19/England/ALDP-A8727A/2020\|EPI_ISL_606084\|20 | .................... | .................... | ..................... | .................... | .................... | .................... |
| O | 3355 hCoV-19/England/ALDP-A87D6A/2020\|EPI_ISL_606086\|20 | .................... | .................... | ..................... | .................... | .................... | .................... |
| O | 3356 hCoV-19/England/ALDP-A87BBB/2020\|EPI_ISL_606087\|20 | .................... | .................... | ..................... | .................... | .................... | .................... |
| O | 3357 hCoV-19/England/ALDP-A86624/2020\|EPI_ISL_606089\|20 | .................... | .................... | ..................... | .................... | .................... | .................... |
| O | 3358 hCoV-19/England/ALDP-A8741A/2020\|EPI_ISL_606090\|20 | .................... | .................... | ..................... | .................... | .................... | .................... |
| O | 3359 hCoV-19/England/ALDP-A862B7/2020\|EPI_ISL_606091\|20 | .................... | .................... | ..................... | .................... | .................... | .................... |
| O | 3360 hCoV-19/England/ALDP-A87386/2020\|EPI_ISL_606092\|20 | .................... | .................... | ..................... | .................... | .................... | .................... |
| O | 3361 hCoV-19/England/ALDP-A87544/2020\|EPI_ISL_606094\|20 | .................... | .................... | ..................... | .................... | .................... | .................... |
| O | 3362 hCoV-19/England/ALDP-A879FD/2020\|EPI_ISL_606095\|20 | .................... | .................... | ..................... | .................... | .................... | .................... |
| O | 3363 hCoV-19/England/ALDP-A86387/2020\|EPI_ISL_606096\|20 | .................... | .................... | ..................... | .................... | .................... | .................... |
| O | 3364 hCoV-19/England/ALDP-A87289/2020\|EPI_ISL_606097\|20 | .................... | .................... | ..................... | .................... | .................... | ..A................. |
| O | 3365 hCoV-19/England/ALDP-A86FCF/2020\|EPI_ISL_606098\|20 | .................... | .................... | ..................... | .................... | .................... | .................... |
| O | 3366 hCoV-19/England/ALDP-A86396/2020\|EPI_ISL_606099\|20 | .................... | .................... | ..................... | .................... | .................... | .................... |
| O | 3367 hCoV-19/England/MILK-A7BFDC/2020\|EPI_ISL_607300\|20 | .................... | .................... | ..................... | .................... | .................... | .................... |
| O | 3368 hCoV-19/England/MILK-A7B947/2020\|EPI_ISL_607301\|20 | .................... | .................... | ..................... | .................... | .................... | .................... |
| O | 3369 hCoV-19/England/MILK-A7B4BF/2020\|EPI_ISL_607302\|20 | .................... | .................... | ..................... | .................... | .................... | .................... |
| O | 3370 hCoV-19/England/MILK-A7BE2A/2020\|EPI_ISL_607304\|20 | .................... | .................... | ..................... | .................... | .................... | .................... |
| O | 3371 hCoV-19/England/MILK-A7B376/2020\|EPI_ISL_607305\|20 | .................... | .................... | ..................... | .................... | .................... | .................... |
| O | 3372 hCoV-19/England/MILK-A7B5DA/2020\|EPI_ISL_607306\|20 | .................... | .................... | ..................... | .................... | .................... | .................... |
| O | 3373 hCoV-19/England/MILK-A7B3A3/2020\|EPI_ISL_607307\|20 | .................... | .................... | ..................... | .................... | .................... | .................... |
| O | 3374 hCoV-19/England/MILK-A7C9BF/2020\|EPI_ISL_607309\|20 | .................... | .................... | ..................... | .................... | .................... | .................... |
| O | 3375 hCoV-19/England/MILK-A7C524/2020\|EPI_ISL_607340\|20 | .................... | .................... | ..................... | .................... | .................... | .................... |
| O | 3376 hCoV-19/Wales/MILK-A7B965/2020\|EPI_ISL_607341\|2020 | .................... | .................... | ..................... | .................... | .................... | .................... |
| O | 3377 hCoV-19/England/MILK-A7B65F/2020\|EPI_ISL_607342\|20 | .................... | .................... | ..................... | .................... | .................... | .................... |

O 3378 hCoV-19/England/MILK-A7C560/2020|EPI_ISL_607390|20 ···················· ···················· ···················· ···················· ···················· ····················
O 3379 hCoV-19/England/MILK-A7BEEE/2020|EPI_ISL_607391|20 ···················· ···················· ···················· ···················· ···················· ····················
O 3380 hCoV-19/England/MILK-A7CB13/2020|EPI_ISL_607392|20 ···················· ···················· ···················· ···················· ···················· ····················
O 3381 hCoV-19/England/MILK-A7B437/2020|EPI_ISL_607393|20 ···················· ···················· ···················· ···················· ···················· ····················
O 3382 hCoV-19/England/MILK-A7BFAF/2020|EPI_ISL_607394|20 ···················· ···················· ···················· ···················· ···················· ····················
O 3383 hCoV-19/England/MILK-A7BB23/2020|EPI_ISL_607395|20 ···················· ·A·················· ···················· ···················· ···················· ··A·················
O 3384 hCoV-19/England/MILK-A7B051/2020|EPI_ISL_607396|20 ···················· ···················· ···················· ···················· ·····T·············· ····················
O 3385 hCoV-19/England/MILK-A7B7C5/2020|EPI_ISL_607398|20 ···················· ···················· ···················· ···················· ···················· ····················
O 3386 hCoV-19/England/MILK-A7BBC9/2020|EPI_ISL_607399|20 ···················· ···················· ···················· ···················· ···················· ····················
O 3387 hCoV-19/England/CAMC-A654F4/2020|EPI_ISL_608560|20 ···················· ···················· ···················· ···················· ···················· ····················
O 3388 hCoV-19/England/ALDP-A68233/2020|EPI_ISL_608600|20 ···················· ···················· ···················· ···················· ···················· ····················
O 3389 hCoV-19/England/ALDP-A68B83/2020|EPI_ISL_608601|20 ···················· ···················· ···················· ···················· ···················· ····················
O 3390 hCoV-19/England/CAMC-A64BC3/2020|EPI_ISL_608602|20 ···················· ···················· ···················· ···················· ···················· ····················
O 3391 hCoV-19/England/ALDP-A67B75/2020|EPI_ISL_608603|20 ···················· ·A·················· ···················· ···················· ···················· ····················
O 3392 hCoV-19/England/CAMC-A64EBB/2020|EPI_ISL_608604|20 ···················· ···················· ···················· ···················· ·····T·············· ····················
O 3393 hCoV-19/England/ALDP-A6838B/2020|EPI_ISL_608605|20 ···················· ···················· ···················· ···················· ···················· ····················
O 3394 hCoV-19/England/ALDP-A68B56/2020|EPI_ISL_608606|20 ···················· ···················· ···················· ···················· ···················· ····················
O 3395 hCoV-19/England/CAMC-A65324/2020|EPI_ISL_608607|20 ···················· ···················· ···················· ···················· ···················· ····················
O 3396 hCoV-19/England/CAMC-A64FD6/2020|EPI_ISL_608608|20 ···················· ···················· ···················· ···················· ···················· ····················
O 3397 hCoV-19/England/ALDP-A68C9F/2020|EPI_ISL_608610|20 ···················· ···················· ···················· ···················· ···················· ····················
O 3398 hCoV-19/England/CAMC-A64CEE/2020|EPI_ISL_608611|20 ···················· ···················· ···················· ···················· ···················· ····················
O 3399 hCoV-19/England/CAMC-A64FB8/2020|EPI_ISL_608612|20 ···················· ···················· ···················· ···················· ···················· ····················
O 3400 hCoV-19/England/CAMC-A65306/2020|EPI_ISL_608613|20 ···················· ···················· ···················· ···················· ···················· ····················
O 3401 hCoV-19/England/ALDP-A67F4C/2020|EPI_ISL_608614|20 ···················· ···················· ···················· ···················· ···················· ····················
O 3402 hCoV-19/England/CAMC-A65157/2020|EPI_ISL_608615|20 ···················· ···················· ···················· ···················· ···················· ····················
O 3403 hCoV-19/England/CAMC-A64B3C/2020|EPI_ISL_608616|20 ···················· ·A·················· ···················· ···················· ···················· ····················
O 3404 hCoV-19/England/ALDP-A68576/2020|EPI_ISL_608617|20 ···················· ···················· ···················· ···················· ···················· ····················
O 3405 hCoV-19/England/CAMC-A64F30/2020|EPI_ISL_608618|20 ···················· ···················· ···················· ···················· ···················· ····················
O 3406 hCoV-19/England/CAMC-A64B4B/2020|EPI_ISL_608619|20 ···················· ···················· ···················· ···················· ···················· ····················
O 3407 hCoV-19/England/ALDP-A67BFD/2020|EPI_ISL_608621|20 ···················· ···················· ···················· ···················· ···················· ····················
O 3408 hCoV-19/Wales/ALDP-A6893E/2020|EPI_ISL_608622|2020 ···················· ···················· ···················· ···················· ···················· ····················
O 3409 hCoV-19/England/ALDP-A6846A/2020|EPI_ISL_608623|20 ···················· ···M················ ···················· ···················· ···················· ····················
O 3410 hCoV-19/England/CAMC-A64BB4/2020|EPI_ISL_608624|20 ···················· ···················· ···················· ···················· ···················· ····················
O 3411 hCoV-19/England/ALDP-A68549/2020|EPI_ISL_608625|20 ···················· ···················· ···················· ···················· ···················· ····················
O 3412 hCoV-19/England/CAMC-A65A5C/2020|EPI_ISL_608626|20 ···················· ···················· ···················· ···················· ···················· ····················
O 3413 hCoV-19/England/CAMC-A653BB/2020|EPI_ISL_608627|20 ···················· ···················· ···················· ···················· ···················· ····················
O 3414 hCoV-19/England/CAMC-A64EE8/2020|EPI_ISL_608628|20 ···················· ···················· ···················· ···················· ···················· ····················
O 3415 hCoV-19/England/CAMC-A650F0/2020|EPI_ISL_608629|20 ···················· ···················· ···················· ···················· ···················· ····················
O 3416 hCoV-19/England/ALDP-A684D3/2020|EPI_ISL_608630|20 ···················· ···················· ···················· ···················· ···················· ····················
O 3417 hCoV-19/England/CAMC-A651EE/2020|EPI_ISL_608631|20 ···················· ···················· ···················· ···················· ···················· ····················
O 3418 hCoV-19/England/CAMC-A650E1/2020|EPI_ISL_608632|20 ···················· ···················· ···················· ···················· ···················· ····················
O 3419 hCoV-19/England/ALDP-A6841F/2020|EPI_ISL_608634|20 ···················· ···················· ···················· ···················· ···················· ····················
O 3420 hCoV-19/England/ALDP-A67F6A/2020|EPI_ISL_608635|20 ···················· ···················· ···················· ···················· ···················· ····················
O 3421 hCoV-19/England/CAMC-A64DFA/2020|EPI_ISL_608636|20 ···················· ···················· ···················· ···················· ···················· ····················
O 3422 hCoV-19/England/CAMC-A64DBE/2020|EPI_ISL_608637|20 ···················· ···················· ···················· ···················· ···················· ····················
O 3423 hCoV-19/England/CAMC-A64BD2/2020|EPI_ISL_608638|20 ···················· ···················· ···················· ···················· ···················· ····················
O 3424 hCoV-19/England/ALDP-A69117/2020|EPI_ISL_608639|20 ···················· ···················· ···················· ···················· ···················· ····················
O 3425 hCoV-19/England/CAMC-A648DB/2020|EPI_ISL_608671|20 ···················· ···················· ···················· ···················· ···················· ····················
O 3426 hCoV-19/England/ALDP-A681AF/2020|EPI_ISL_608672|20 ···················· ···················· ···················· ···················· ···················· ····················
O 3427 hCoV-19/England/CAMC-A6589E/2020|EPI_ISL_608673|20 ···················· ···················· ···················· ···················· ·····T·············· ····················
O 3428 hCoV-19/England/ALDP-A67F79/2020|EPI_ISL_608674|20 ···················· ···················· ···················· ···················· ···················· ····················
O 3429 hCoV-19/England/204221328/2020|EPI_ISL_609890|2020 ···················· ···················· ···················· ···················· ···················· ····················
O 3430 hCoV-19/England/204240010/2020|EPI_ISL_609891|2020 ···················· ···················· ···················· ···················· ···················· ····················
O 3431 hCoV-19/England/204260476/2020|EPI_ISL_609901|2020 ···················· ···················· ···················· ···················· ···················· ····················
O 3432 hCoV-19/England/204380928/2020|EPI_ISL_609902|2020 ···················· ···················· ···················· ···················· ···················· ····················
O 3433 hCoV-19/England/204380929/2020|EPI_ISL_609903|2020 ···················· ···················· ···················· ···················· ···················· ····················
O 3434 hCoV-19/England/204380930/2020|EPI_ISL_609904|2020 ···················· ···················· ···················· ···················· ···················· ····················
O 3435 hCoV-19/England/204380931/2020|EPI_ISL_609905|2020 ···················· ···················· ···················· ···················· ···················· ····················
O 3436 hCoV-19/England/204380932/2020|EPI_ISL_609906|2020 ···················· ···················· ···················· ···················· ···················· ····················
O 3437 hCoV-19/England/204380933/2020|EPI_ISL_609907|2020 ···················· ···················· ···················· ···················· ···················· ····················
O 3438 hCoV-19/England/204380934/2020|EPI_ISL_609908|2020 ···················· ···················· ···················· ···················· ···················· ····················
O 3439 hCoV-19/England/CAMC-A81261/2020|EPI_ISL_610880|20 ···················· ···················· ···················· ···················· ···················· ····················
O 3440 hCoV-19/England/CAMC-A8197B/2020|EPI_ISL_610882|20 ···················· ···················· ···················· ···················· ···················· ····················
O 3441 hCoV-19/England/CAMC-A812E9/2020|EPI_ISL_610900|20 ···················· ···················· ···················· ···················· ···················· ····················
O 3442 hCoV-19/England/CAMC-A82190/2020|EPI_ISL_610901|20 ···················· ·A·················· ···················· ···················· ···················· ····················
O 3443 hCoV-19/England/CAMC-A81AF0/2020|EPI_ISL_610902|20 ···················· ···················· ···················· ···················· ···················· ····················
O 3444 hCoV-19/England/CAMC-A81B93/2020|EPI_ISL_610903|20 ···················· ···················· ···················· ···················· ···················· ····················
O 3445 hCoV-19/England/CAMC-A82EB7/2020|EPI_ISL_610904|20 ···················· ···················· ···················· ···················· ···················· ····················
O 3446 hCoV-19/England/CAMC-A8238B/2020|EPI_ISL_610905|20 ···················· ···················· ···················· ···················· ···················· ····················
O 3447 hCoV-19/England/CAMC-A81CCD/2020|EPI_ISL_610906|20 ···················· ···················· ···················· ···················· ···················· ····················
O 3448 hCoV-19/England/CAMC-A8250D/2020|EPI_ISL_610907|20 ···················· ···················· ···················· ···················· ···················· ····················
O 3449 hCoV-19/England/CAMC-A81216/2020|EPI_ISL_610908|20 ···················· ···················· ···················· ···················· ···················· ····················
O 3450 hCoV-19/England/CAMC-A82DD8/2020|EPI_ISL_610909|20 ···················· ···················· ···················· ···················· ···················· ····················
O 3451 hCoV-19/England/CAMC-A81D7F/2020|EPI_ISL_610910|20 ···················· ···················· ···················· ···················· ···················· ····················
O 3452 hCoV-19/England/CAMC-A81BC0/2020|EPI_ISL_610911|20 ···················· ·A·················· ···················· ···················· ···················· ····················
O 3453 hCoV-19/England/CAMC-A80D70/2020|EPI_ISL_610912|20 ···················· ···················· ···················· ···················· ···················· ····················
O 3454 hCoV-19/England/CAMC-A829A7/2020|EPI_ISL_610915|20 ···················· ···················· ···················· ···················· ···················· ····················
O 3455 hCoV-19/England/CAMC-A8295C/2020|EPI_ISL_610916|20 ···················· ···················· ···················· ···················· ···················· ····················
O 3456 hCoV-19/England/CAMC-A81DD9/2020|EPI_ISL_610917|20 ···················· ···················· ···················· ···················· ···················· ····················
O 3457 hCoV-19/England/CAMC-A829F2/2020|EPI_ISL_610920|20 ···················· ···················· ···················· ···················· ···················· ····················

| | | | | | | | |
|---|---|---|---|---|---|---|---|
| O | 3458 hCoV-19/England/CAMC-A83047/2020\|EPI_ISL_610921\|20 | ···················· | ···················· | ····················· | ···················· | ······T············· | ···················· |
| O | 3459 hCoV-19/England/ALDP-A93633/2020\|EPI_ISL_610922\|20 | ···················· | ···················· | ····················· | ···················· | ···················· | ···················· |
| O | 3460 hCoV-19/England/ALDP-A92BDB/2020\|EPI_ISL_610923\|20 | ···················· | ···················· | ····················· | ···················· | ···················· | ···················· |
| O | 3461 hCoV-19/England/ALDP-A927E6/2020\|EPI_ISL_610924\|20 | ···················· | ···················· | ····················· | ···················· | ···················· | ···················· |
| O | 3462 hCoV-19/England/ALDP-A93A55/2020\|EPI_ISL_610925\|20 | ···················· | ···················· | ····················· | ···················· | ···················· | ···················· |
| O | 3463 hCoV-19/England/ALDP-A9287A/2020\|EPI_ISL_610926\|20 | ···················· | ···················· | ····················· | ···················· | ···················· | ···················· |
| O | 3464 hCoV-19/England/ALDP-A9390D/2020\|EPI_ISL_610927\|20 | ···················· | ·········T·········· | ····················· | ···················· | ···················· | ···················· |
| O | 3465 hCoV-19/England/ALDP-A943FF/2020\|EPI_ISL_610931\|20 | ···················· | ···················· | ····················· | ···················· | ···················· | ···················· |
| O | 3466 hCoV-19/England/ALDP-A93949/2020\|EPI_ISL_610933\|20 | ···················· | ···················· | ····················· | ···················· | ···················· | ···················· |
| O | 3467 hCoV-19/England/ALDP-A92C14/2020\|EPI_ISL_610934\|20 | ···················· | ···················· | ····················· | ···················· | ···················· | ···················· |
| O | 3468 hCoV-19/England/ALDP-A93D01/2020\|EPI_ISL_610936\|20 | ···················· | ···················· | ····················· | ···················· | ···················· | ···················· |
| O | 3469 hCoV-19/England/ALDP-A93A37/2020\|EPI_ISL_610937\|20 | ···················· | ···················· | ····················· | ···················· | ···················· | ···················· |
| O | 3470 hCoV-19/England/ALDP-A928E3/2020\|EPI_ISL_610938\|20 | ···················· | ···················· | ····················· | ···················· | ···················· | ···················· |
| O | 3471 hCoV-19/England/ALDP-A93879/2020\|EPI_ISL_610939\|20 | ···················· | ···················· | ····················· | ···················· | ···················· | ···················· |
| O | 3472 hCoV-19/England/ALDP-A92722/2020\|EPI_ISL_610940\|20 | ···················· | ···················· | ····················· | ···················· | ···················· | ···················· |
| O | 3473 hCoV-19/England/ALDP-A925EC/2020\|EPI_ISL_610941\|20 | ···················· | ···················· | ····················· | ···················· | ···················· | ···················· |
| O | 3474 hCoV-19/England/ALDP-A94429/2020\|EPI_ISL_610942\|20 | ···················· | ···················· | ····················· | ···················· | ···················· | ···················· |
| O | 3475 hCoV-19/England/ALDP-A9241C/2020\|EPI_ISL_610943\|20 | ···················· | ···················· | ····················· | ···················· | ···················· | ···················· |
| O | 3476 hCoV-19/England/ALDP-A92EC3/2020\|EPI_ISL_610944\|20 | ···················· | ···················· | ····················· | ···················· | ···················· | ···················· |
| O | 3477 hCoV-19/England/ALDP-A93F38/2020\|EPI_ISL_610945\|20 | ···················· | ···················· | ····················· | ···················· | ···················· | ···················· |
| O | 3478 hCoV-19/England/ALDP-A92DD5/2020\|EPI_ISL_610946\|20 | ···················· | ···················· | ····················· | ···················· | ···················· | ···················· |
| O | 3479 hCoV-19/England/ALDP-A92E0F/2020\|EPI_ISL_610947\|20 | ···················· | ···················· | ····················· | ···················· | ···················· | ···················· |
| O | 3480 hCoV-19/England/ALDP-A93703/2020\|EPI_ISL_610948\|20 | ···················· | ···················· | ····················· | ···················· | ···················· | ···················· |
| O | 3481 hCoV-19/England/ALDP-A9268F/2020\|EPI_ISL_610949\|20 | ···················· | ···················· | ····················· | ···················· | ···················· | ···················· |
| O | 3482 hCoV-19/England/ALDP-A93AA0/2020\|EPI_ISL_610950\|20 | ···················· | ···················· | ····················· | ···················· | ···················· | ···················· |
| O | 3483 hCoV-19/England/ALDP-A93B07/2020\|EPI_ISL_610951\|20 | ···················· | ···················· | ····················· | ···················· | ···················· | ···················· |
| O | 3484 hCoV-19/England/ALDP-A92616/2020\|EPI_ISL_610952\|20 | ···················· | ···················· | ····················· | ···················· | ···················· | ···················· |
| O | 3486 hCoV-19/Wales/PHWC-48172C/2020\|EPI_ISL_613101\|2020 | ···················· | ···················· | ····················· | ···················· | ···················· | ···················· |
| O | 3487 hCoV-19/Wales/PHWC-48173B/2020\|EPI_ISL_613102\|2020 | ···················· | ···················· | ····················· | ···················· | ···················· | ···················· |
| O | 3490 hCoV-19/Wales/PHWC-481768/2020\|EPI_ISL_613105\|2020 | ···················· | ···················· | ····················· | ···················· | ···················· | ···················· |
| O | 3491 hCoV-19/Wales/PHWC-481786/2020\|EPI_ISL_613106\|2020 | ···················· | ···················· | ····················· | ···················· | ···················· | ···················· |
| O | 3493 hCoV-19/Wales/PHWC-4817A4/2020\|EPI_ISL_613108\|2020 | ···················· | ···················· | ····················· | ···················· | ···················· | ···················· |
| O | 3494 hCoV-19/Wales/PHWC-4817B3/2020\|EPI_ISL_613109\|2020 | ···················· | ···················· | ····················· | ···················· | ···················· | ···················· |
| O | 3495 hCoV-19/Wales/PHWC-4817C2/2020\|EPI_ISL_613110\|2020 | ···················· | ···················· | ····················· | ···················· | ···················· | ···················· |
| O | 3496 hCoV-19/Wales/PHWC-4817D1/2020\|EPI_ISL_613111\|2020 | ···················· | ···················· | ····················· | ···················· | ···················· | ···················· |
| O | 3497 hCoV-19/Wales/PHWC-4817E0/2020\|EPI_ISL_613112\|2020 | ···················· | ···················· | ····················· | ···················· | ···················· | ···················· |
| O | 3498 hCoV-19/Wales/PHWC-4817FF/2020\|EPI_ISL_613113\|2020 | ···················· | ···················· | ····················· | ···················· | ···················· | ···················· |
| O | 3499 hCoV-19/Wales/PHWC-48180B/2020\|EPI_ISL_613114\|2020 | ···················· | ···················· | ····················· | ···················· | ···················· | ···················· |
| O | 3500 hCoV-19/Wales/PHWC-48181A/2020\|EPI_ISL_613115\|2020 | ···················· | ···················· | ····················· | ···················· | ···················· | ···················· |
| O | 3503 hCoV-19/Wales/PHWC-481856/2020\|EPI_ISL_613118\|2020 | ···················· | ···················· | ····················· | ···················· | ···················· | ···················· |
| O | 3504 hCoV-19/Wales/PHWC-481865/2020\|EPI_ISL_613119\|2020 | ···················· | ···················· | ····················· | ···················· | ···················· | ···················· |
| O | 3505 hCoV-19/Wales/PHWC-4818A1/2020\|EPI_ISL_613120\|2020 | ···················· | ···················· | ····················· | ···················· | ···················· | ···················· |
| O | 3506 hCoV-19/Wales/PHWC-4818DE/2020\|EPI_ISL_613121\|2020 | ···················· | ···················· | ····················· | ···················· | ···················· | ···················· |
| O | 3507 hCoV-19/Wales/PHWC-4818ED/2020\|EPI_ISL_613122\|2020 | ···················· | ···················· | ····················· | ···················· | ···················· | ···················· |
| O | 3508 hCoV-19/Wales/PHWC-4818FC/2020\|EPI_ISL_613123\|2020 | ···················· | ···················· | ····················· | ···················· | ···················· | ···················· |
| O | 3509 hCoV-19/Wales/PHWC-481917/2020\|EPI_ISL_613124\|2020 | ···················· | ···················· | ····················· | ···················· | ···················· | ·········N·········· |
| O | 3510 hCoV-19/Wales/PHWC-481926/2020\|EPI_ISL_613125\|2020 | ···················· | ···················· | ····················· | ···················· | ···················· | ···················· |
| O | 3511 hCoV-19/Wales/PHWC-481944/2020\|EPI_ISL_613126\|2020 | ···················· | ···················· | ····················· | ···················· | ···················· | ···················· |
| O | 3512 hCoV-19/Wales/PHWC-481953/2020\|EPI_ISL_613127\|2020 | ···················· | ···················· | ····················· | ···················· | ···················· | ···················· |
| O | 3513 hCoV-19/Wales/PHWC-481971/2020\|EPI_ISL_613128\|2020 | ···················· | ···················· | ····················· | ···················· | ···················· | ···················· |
| O | 3515 hCoV-19/Wales/PHWC-48199F/2020\|EPI_ISL_613130\|2020 | ···················· | ···················· | ····················· | ···················· | ···················· | ···················· |
| O | 3516 hCoV-19/Wales/PHWC-4819BD/2020\|EPI_ISL_613131\|2020 | ···················· | ···················· | ····················· | ···················· | ···················· | ···················· |
| O | 3517 hCoV-19/Wales/PHWC-4819CC/2020\|EPI_ISL_613132\|2020 | ···················· | ···················· | ····················· | ···················· | ···················· | ···················· |
| O | 3518 hCoV-19/Wales/PHWC-4819DB/2020\|EPI_ISL_613133\|2020 | ···················· | ···················· | ····················· | ···················· | ···················· | ···················· |
| O | 3519 hCoV-19/Wales/PHWC-4819EA/2020\|EPI_ISL_613134\|2020 | ···················· | ···················· | ····················· | ···················· | ···················· | ···················· |
| O | 3520 hCoV-19/Wales/PHWC-4819F9/2020\|EPI_ISL_613135\|2020 | ···················· | ···················· | ····················· | ···················· | ···················· | ···················· |
| O | 3521 hCoV-19/Wales/PHWC-481A05/2020\|EPI_ISL_613136\|2020 | ···················· | ···················· | ····················· | ···················· | ···················· | ···················· |
| O | 3522 hCoV-19/Wales/PHWC-481A14/2020\|EPI_ISL_613137\|2020 | ···················· | ···················· | ····················· | ···················· | ···················· | ···················· |
| O | 3523 hCoV-19/Wales/PHWC-481A6F/2020\|EPI_ISL_613138\|2020 | ···················· | ···················· | ····················· | ······G············· | ···················· | ···················· |
| O | 3525 hCoV-19/Wales/PHWC-481AAB/2020\|EPI_ISL_613140\|2020 | ···················· | ···················· | ····················· | ···················· | ···················· | ···················· |
| O | 3527 hCoV-19/Wales/PHWC-481AC9/2020\|EPI_ISL_613142\|2020 | ···················· | ···················· | ····················· | ···················· | ···················· | ···················· |
| O | 3528 hCoV-19/Wales/PHWC-481AD8/2020\|EPI_ISL_613143\|2020 | ···················· | ···················· | ····················· | ···················· | ···················· | ···················· |
| O | 3530 hCoV-19/Wales/PHWC-481AF6/2020\|EPI_ISL_613145\|2020 | ···················· | ···················· | ····················· | ···················· | ···················· | ···················· |
| O | 3533 hCoV-19/Wales/PHWC-481B6C/2020\|EPI_ISL_613148\|2020 | ···················· | ···················· | ····················· | ···················· | ···················· | ···················· |
| O | 3534 hCoV-19/Wales/PHWC-481B7B/2020\|EPI_ISL_613149\|2020 | ···················· | ···················· | ····················· | ···················· | ···················· | ···················· |
| O | 3537 hCoV-19/Wales/PHWC-481BA8/2020\|EPI_ISL_613152\|2020 | ···················· | ···················· | ····················· | ···················· | ···················· | ···················· |
| O | 3538 hCoV-19/Wales/PHWC-481BC6/2020\|EPI_ISL_613153\|2020 | ···················· | ···················· | ····················· | ···················· | ···················· | ···················· |
| O | 3539 hCoV-19/Wales/PHWC-481BF3/2020\|EPI_ISL_613154\|2020 | ···················· | ···················· | ····················· | ···················· | ···················· | ···················· |
| O | 3540 hCoV-19/Wales/PHWC-481C0F/2020\|EPI_ISL_613155\|2020 | ···················· | ···················· | ····················· | ···················· | ···················· | ···················· |
| O | 3541 hCoV-19/Wales/PHWC-481C1E/2020\|EPI_ISL_613156\|2020 | ···················· | ···················· | ····················· | ···················· | ···················· | ···················· |
| O | 3542 hCoV-19/Wales/PHWC-481C2D/2020\|EPI_ISL_613157\|2020 | ···················· | ···················· | ····················· | ···················· | ···················· | ···················· |
| O | 3543 hCoV-19/Wales/PHWC-481C3C/2020\|EPI_ISL_613158\|2020 | ···················· | ···················· | ····················· | ···················· | ···················· | ···················· |
| O | 3545 hCoV-19/Wales/PHWC-481C5A/2020\|EPI_ISL_613160\|2020 | ···················· | ···················· | ····················· | ···················· | ···················· | ···················· |
| O | 3546 hCoV-19/Wales/PHWC-481C78/2020\|EPI_ISL_613161\|2020 | ···················· | ···················· | ····················· | ···················· | ···················· | ···················· |
| O | 3547 hCoV-19/Wales/PHWC-481CA5/2020\|EPI_ISL_613162\|2020 | ···················· | ···················· | ····················· | ···················· | ···················· | ···················· |
| O | 3549 hCoV-19/Wales/PHWC-481CF0/2020\|EPI_ISL_613164\|2020 | ···················· | ···················· | ····················· | ···················· | ···················· | ···················· |
| O | 3550 hCoV-19/Wales/PHWC-481D0C/2020\|EPI_ISL_613165\|2020 | ···················· | ···················· | ····················· | ···················· | ···················· | ···················· |
| O | 3551 hCoV-19/Wales/PHWC-481D2A/2020\|EPI_ISL_613166\|2020 | ···················· | ···················· | ····················· | ···················· | ···················· | ···················· |
| O | 3552 hCoV-19/Wales/PHWC-481D57/2020\|EPI_ISL_613167\|2020 | ···················· | ···················· | ····················· | ···················· | ···················· | ···················· |
| O | 3553 hCoV-19/Wales/PHWC-481D66/2020\|EPI_ISL_613168\|2020 | ···················· | ···················· | ····················· | ···················· | ···················· | ···················· |

| | | | | | | | |
|---|---|---|---|---|---|---|---|
| O | 3554 hCoV-19/Wales/PHWC-481D75/2020\|EPI_ISL_613169\|2020 | ···················· | ···················· | ···················· | ···················· | ···················· | ···················· |
| O | 3555 hCoV-19/Wales/PHWC-481DC0/2020\|EPI_ISL_613170\|2020 | ···················· | ···················· | ···················· | ···················· | ···················· | ···················· |
| O | 3556 hCoV-19/Wales/PHWC-481DDF/2020\|EPI_ISL_613171\|2020 | ···················· | ···················· | ···················· | ···················· | ···················· | ···················· |
| O | 3557 hCoV-19/Wales/PHWC-481DEE/2020\|EPI_ISL_613172\|2020 | ···················· | ···················· | ···················· | ···················· | ···················· | ···················· |
| O | 3558 hCoV-19/Wales/PHWC-481DFD/2020\|EPI_ISL_613173\|2020 | ···················· | ···················· | ···················· | ···················· | ···················· | ···················· |
| O | 3559 hCoV-19/Wales/PHWC-481E09/2020\|EPI_ISL_613174\|2020 | ···················· | ···················· | ···················· | ···················· | ···················· | ···················· |
| O | 3560 hCoV-19/Wales/PHWC-481E18/2020\|EPI_ISL_613175\|2020 | ···················· | ···················· | ···················· | ···················· | ···················· | ···················· |
| O | 3561 hCoV-19/Wales/PHWC-481E27/2020\|EPI_ISL_613176\|2020 | ···················· | ···················· | ···················· | ···················· | ···················· | ···················· |
| O | 3562 hCoV-19/Wales/PHWC-481E36/2020\|EPI_ISL_613177\|2020 | ···················· | ···················· | ···················· | ···················· | ···················· | ···················· |
| O | 3563 hCoV-19/Wales/PHWC-481E45/2020\|EPI_ISL_613178\|2020 | ···················· | ···················· | ···················· | ···················· | ···················· | ···················· |
| O | 3564 hCoV-19/Wales/PHWC-481E54/2020\|EPI_ISL_613179\|2020 | ···················· | ···················· | ···················· | ···················· | ···················· | ···················· |
| O | 3565 hCoV-19/Wales/PHWC-481E63/2020\|EPI_ISL_613180\|2020 | ···················· | ···················· | ···················· | ···················· | ···················· | ···················· |
| O | 3567 hCoV-19/Wales/PHWC-481E81/2020\|EPI_ISL_613182\|2020 | ···················· | ···················· | ···················· | ···················· | ···················· | ···················· |
| O | 3568 hCoV-19/Wales/PHWC-481EAF/2020\|EPI_ISL_613183\|2020 | ···················· | ···················· | ···················· | ···················· | ···················· | ···················· |
| O | 3569 hCoV-19/Wales/PHWC-481F06/2020\|EPI_ISL_613184\|2020 | ···················· | ···················· | ···················· | ···················· | ···················· | ···················· |
| O | 3570 hCoV-19/Wales/PHWC-481F15/2020\|EPI_ISL_613185\|2020 | ···················· | ···················· | ···················· | ···················· | ···················· | ···················· |
| O | 3571 hCoV-19/Wales/PHWC-481F24/2020\|EPI_ISL_613186\|2020 | ···················· | ···················· | ···················· | ···················· | ···················· | ···················· |
| O | 3572 hCoV-19/Wales/PHWC-481F33/2020\|EPI_ISL_613187\|2020 | ···················· | ···················· | ···················· | ···················· | ···················· | ···················· |
| O | 3573 hCoV-19/Wales/PHWC-481F51/2020\|EPI_ISL_613188\|2020 | ···················· | ···················· | ···················· | ···················· | ···················· | ···················· |
| O | 3574 hCoV-19/Wales/PHWC-481F7F/2020\|EPI_ISL_613189\|2020 | ···················· | ···················· | ···················· | ···················· | ···················· | ···················· |
| O | 3575 hCoV-19/Wales/PHWC-481F9D/2020\|EPI_ISL_613190\|2020 | ···················· | ···················· | ···················· | ···················· | ···················· | ···················· |
| O | 3576 hCoV-19/Wales/PHWC-481FD9/2020\|EPI_ISL_613191\|2020 | ···················· | ···················· | ···················· | ···················· | ···················· | ···················· |
| O | 3578 hCoV-19/Wales/PHWC-482011/2020\|EPI_ISL_613193\|2020 | ···················· | ···················· | ···················· | ···················· | ···················· | ···················· |
| O | 3579 hCoV-19/Wales/PHWC-48203F/2020\|EPI_ISL_613194\|2020 | ···················· | ···················· | ···················· | ···················· | ···················· | ···················· |
| O | 3580 hCoV-19/Wales/PHWC-48204E/2020\|EPI_ISL_613195\|2020 | ···················· | ···················· | ···················· | ···················· | ···················· | ···················· |
| O | 3581 hCoV-19/Wales/PHWC-48205D/2020\|EPI_ISL_613196\|2020 | ···················· | ···················· | ···················· | ···················· | ···················· | ···················· |
| O | 3582 hCoV-19/Wales/PHWC-48206C/2020\|EPI_ISL_613197\|2020 | ···················· | ···················· | ···················· | ···················· | ···················· | ···················· |
| O | 3583 hCoV-19/Wales/PHWC-4820C6/2020\|EPI_ISL_613198\|2020 | ···················· | ···················· | ···················· | ···················· | ···················· | ···················· |
| O | 3584 hCoV-19/Wales/PHWC-48210F/2020\|EPI_ISL_613199\|2020 | ···················· | ···················· | ···················· | ···················· | ···················· | ···················· |
| O | 3585 hCoV-19/England/ALDP-A6CABF/2020\|EPI_ISL_607201\|20 | ···················· | ···················· | ···················· | ···················· | ···················· | ···················· |
| O | 3586 hCoV-19/England/ALDP-A6FC7A/2020\|EPI_ISL_607202\|20 | ···················· | ···················· | ···················· | ···················· | ···················· | ···················· |
| O | 3587 hCoV-19/England/ALDP-A6DE3A/2020\|EPI_ISL_607203\|20 | ···················· | ···················· | ···················· | ···················· | ···················· | ···················· |
| O | 3588 hCoV-19/England/ALDP-A6F7C4/2020\|EPI_ISL_607204\|20 | ···················· | ···················· | ···················· | ···················· | ···················· | ···················· |
| O | 3589 hCoV-19/England/ALDP-A6C7B8/2020\|EPI_ISL_607205\|20 | ···················· | ···················· | ···················· | ···················· | ···················· | ···················· |
| O | 3590 hCoV-19/England/ALDP-A6EC99/2020\|EPI_ISL_607206\|20 | ···················· | ···················· | ···················· | ···················· | ···················· | ···················· |
| O | 3591 hCoV-19/England/ALDP-A6EC8A/2020\|EPI_ISL_607207\|20 | ···················· | ···················· | ···················· | ···················· | ···················· | ···················· |
| O | 3592 hCoV-19/England/ALDP-A6FBF5/2020\|EPI_ISL_607208\|20 | ···················· | ···················· | ···················· | ···················· | ···················· | ···················· |
| O | 3593 hCoV-19/England/ALDP-A6CC13/2020\|EPI_ISL_607209\|20 | ···················· | ···················· | ···················· | ···················· | ···················· | ···················· |
| O | 3594 hCoV-19/England/ALDP-A6D9EE/2020\|EPI_ISL_607210\|20 | ···················· | ···················· | ···················· | ···················· | ···················· | ···················· |
| O | 3595 hCoV-19/England/ALDP-A6D67E/2020\|EPI_ISL_607211\|20 | ···················· | ···················· | ···················· | ···················· | ···················· | ···················· |
| O | 3596 hCoV-19/England/ALDP-A6FC01/2020\|EPI_ISL_607212\|20 | ···················· | ···················· | ···················· | ···················· | ···················· | ···················· |
| O | 3597 hCoV-19/England/ALDP-A6EDF0/2020\|EPI_ISL_607213\|20 | ···················· | ···················· | ···················· | ···················· | ···················· | ···················· |
| O | 3598 hCoV-19/England/ALDP-A6D975/2020\|EPI_ISL_607214\|20 | ···················· | ···················· | ···················· | ···················· | ···················· | ···················· |
| O | 3599 hCoV-19/England/ALDP-A6E516/2020\|EPI_ISL_607215\|20 | ···················· | ·············C······ | ···················· | ···················· | ···················· | ···················· |
| O | 3600 hCoV-19/England/ALDP-A6E6C8/2020\|EPI_ISL_607217\|20 | ···················· | ···················· | ···················· | ···················· | ···················· | ···················· |
| O | 3601 hCoV-19/England/ALDP-A6F375/2020\|EPI_ISL_607219\|20 | ···················· | ···················· | ···················· | ···················· | ···················· | ···················· |
| O | 3602 hCoV-19/England/ALDP-A6EEB1/2020\|EPI_ISL_607220\|20 | ···················· | ···················· | ···················· | ···················· | ···················· | ···················· |
| O | 3603 hCoV-19/England/ALDP-A6F946/2020\|EPI_ISL_607221\|20 | ···················· | ···················· | ···················· | ···················· | ···················· | ···················· |
| O | 3604 hCoV-19/England/ALDP-A6E358/2020\|EPI_ISL_607222\|20 | ···················· | ···················· | ···················· | ···················· | ···················· | ···················· |
| O | 3605 hCoV-19/England/ALDP-A6E32B/2020\|EPI_ISL_607224\|20 | ···················· | ···················· | ···················· | ···················· | ···················· | ···················· |
| O | 3606 hCoV-19/England/ALDP-A6E69B/2020\|EPI_ISL_607225\|20 | ···················· | ···················· | ···················· | ···················· | ···················· | ···················· |
| O | 3607 hCoV-19/England/ALDP-A6E14F/2020\|EPI_ISL_607226\|20 | ···················· | ···················· | ···················· | ···················· | ···················· | ···················· |
| O | 3608 hCoV-19/England/ALDP-A6D5CC/2020\|EPI_ISL_607227\|20 | ···················· | ···················· | ···················· | ···················· | ···················· | ···················· |
| O | 3609 hCoV-19/England/ALDP-A6F8A3/2020\|EPI_ISL_607228\|20 | ···················· | ···················· | ···················· | ···················· | ···················· | ···················· |
| O | 3610 hCoV-19/England/ALDP-A6FD59/2020\|EPI_ISL_607229\|20 | ···················· | ···················· | ···················· | ···················· | ···················· | ···················· |
| O | 3611 hCoV-19/England/ALDP-A6E613/2020\|EPI_ISL_607230\|20 | ···················· | ···················· | ···················· | ···················· | ···················· | ···················· |
| O | 3612 hCoV-19/England/ALDP-A6F75B/2020\|EPI_ISL_607231\|20 | ···················· | ···················· | ···················· | ···················· | ···················· | ···················· |
| O | 3613 hCoV-19/England/ALDP-A6FDFF/2020\|EPI_ISL_607233\|20 | ···················· | ···················· | ···················· | ···················· | ···················· | ···················· |
| O | 3614 hCoV-19/England/ALDP-A6D5F9/2020\|EPI_ISL_607234\|20 | ···················· | ···················· | ···················· | ···················· | ···················· | ···················· |
| O | 3615 hCoV-19/England/ALDP-A6F21E/2020\|EPI_ISL_607236\|20 | ···················· | ···················· | ···················· | ···················· | ···················· | ···················· |
| O | 3616 hCoV-19/England/ALDP-A6F779/2020\|EPI_ISL_607237\|20 | ···················· | ···················· | ···················· | ···················· | ···················· | ···················· |
| O | 3617 hCoV-19/England/ALDP-A6F83A/2020\|EPI_ISL_607238\|20 | ···················· | ···················· | ···················· | ···················· | ···················· | ···················· |
| O | 3618 hCoV-19/England/ALDP-A6C9D0/2020\|EPI_ISL_607239\|20 | ···················· | ···················· | ···················· | ···················· | ···················· | ···················· |
| O | 3619 hCoV-19/England/ALDP-A6E4FB/2020\|EPI_ISL_607240\|20 | ···················· | ···················· | ···················· | ···················· | ···················· | ···················· |
| O | 3620 hCoV-19/England/ALDP-A6D641/2020\|EPI_ISL_607241\|20 | ···················· | ···················· | ···················· | ···················· | ···················· | ···················· |
| O | 3621 hCoV-19/England/ALDP-A6E534/2020\|EPI_ISL_607243\|20 | ···················· | ···················· | ···················· | ···················· | ····T··············· | ···················· |
| O | 3622 hCoV-19/England/ALDP-A6F885/2020\|EPI_ISL_607244\|20 | ···················· | ···················· | ···················· | ···················· | ···················· | ···················· |
| O | 3623 hCoV-19/England/ALDP-A6CA82/2020\|EPI_ISL_607245\|20 | ···················· | ···················· | ···················· | ···················· | ···················· | ···················· |
| O | 3624 hCoV-19/England/ALDP-A6F858/2020\|EPI_ISL_607247\|20 | ···················· | ···················· | ···················· | ···················· | ···················· | ···················· |
| O | 3625 hCoV-19/England/ALDP-A6FC2F/2020\|EPI_ISL_607248\|20 | ···················· | ···················· | ···················· | ···················· | ···················· | ···················· |
| O | 3626 hCoV-19/England/ALDP-A6ECC6/2020\|EPI_ISL_607249\|20 | ···················· | ···················· | ···················· | ···················· | ···················· | ···················· |
| O | 3627 hCoV-19/England/ALDP-A6E67D/2020\|EPI_ISL_607250\|20 | ···················· | ···················· | ···················· | ···················· | ···················· | ···················· |
| O | 3628 hCoV-19/England/ALDP-A6F8C1/2020\|EPI_ISL_607251\|20 | ···················· | ···················· | ···················· | ···················· | ···················· | ···················· |
| O | 3629 hCoV-19/England/ALDP-A6CA91/2020\|EPI_ISL_607252\|20 | ···················· | ···················· | ···················· | ···················· | ···················· | ···················· |
| O | 3630 hCoV-19/England/ALDP-A6F700/2020\|EPI_ISL_607253\|20 | ···················· | ···················· | ···················· | ···················· | ···················· | ···················· |
| O | 3631 hCoV-19/England/ALDP-A6D632/2020\|EPI_ISL_607254\|20 | ···················· | ···················· | ···················· | ···················· | ···················· | ···················· |
| O | 3632 hCoV-19/England/ALDP-A6D993/2020\|EPI_ISL_607255\|20 | ···················· | ···················· | ···················· | ···················· | ···················· | ···················· |
| O | 3633 hCoV-19/England/ALDP-A6F2E1/2020\|EPI_ISL_607256\|20 | ···················· | ···················· | ···················· | ···················· | ···················· | ···················· |
| O | 3634 hCoV-19/England/ALDP-A6DD88/2020\|EPI_ISL_607258\|20 | ···················· | ···················· | ···················· | ···················· | ···················· | ···················· |
| O | 3635 hCoV-19/England/ALDP-A6EC20/2020\|EPI_ISL_607259\|20 | ···················· | ···················· | ···················· | ···················· | ···················· | ···················· |

| | | | | | | | | |
|---|---|---|---|---|---|---|---|---|
| O | 3636 | hCoV-19/England/ALDP-A6FBC8/2020\|EPI_ISL_607260\|20 | ···················· | ···················· | ····················· | ···················· | ···················· | ···················· |
| O | 3637 | hCoV-19/England/ALDP-A6F964/2020\|EPI_ISL_607261\|20 | ···················· | ···················· | ····················· | ···················· | ···················· | ···················· |
| O | 3638 | hCoV-19/England/ALDP-A6FDD1/2020\|EPI_ISL_607262\|20 | ···················· | ···················· | ····················· | ···················· | ···················· | ···················· |
| O | 3639 | hCoV-19/England/ALDP-A6CA37/2020\|EPI_ISL_607263\|20 | ···················· | ···················· | ····················· | ···················· | ···················· | ···················· |
| O | 3640 | hCoV-19/England/ALDP-A6E394/2020\|EPI_ISL_607264\|20 | ···················· | ···················· | ····················· | ···················· | ···················· | ···················· |
| O | 3641 | hCoV-19/England/ALDP-A6D5BD/2020\|EPI_ISL_607265\|20 | ···················· | ···················· | ····················· | ···················· | ···················· | ···················· |
| O | 3642 | hCoV-19/England/ALDP-A6E6D7/2020\|EPI_ISL_607266\|20 | ···················· | ···················· | ····················· | ···················· | ···················· | ···················· |
| O | 3643 | hCoV-19/England/ALDP-A6FCF2/2020\|EPI_ISL_607267\|20 | ···················· | ···················· | ····················· | ···················· | ···················· | ···················· |
| O | 3644 | hCoV-19/England/ALDP-A6FD77/2020\|EPI_ISL_607268\|20 | ···················· | ···················· | ····················· | ···················· | ···················· | ···················· |
| O | 3645 | hCoV-19/England/ALDP-A6DDB5/2020\|EPI_ISL_607269\|20 | ···················· | ···················· | ····················· | ···················· | ···················· | ···················· |
| O | 3646 | hCoV-19/England/ALDP-A6C721/2020\|EPI_ISL_607270\|20 | ···················· | ···················· | ····················· | ···················· | ···················· | ···················· |
| O | 3647 | hCoV-19/England/ALDP-A6E0D9/2020\|EPI_ISL_607271\|20 | ···················· | ·A·················· | ····················· | ···················· | ···················· | ···················· |
| O | 3648 | hCoV-19/England/ALDP-A6F1F3/2020\|EPI_ISL_607272\|20 | ···················· | ···················· | ····················· | ···················· | ···················· | ···················· |
| O | 3649 | hCoV-19/England/ALDP-A6E0E8/2020\|EPI_ISL_607273\|20 | ···················· | ·A·················· | ····················· | ···················· | ···················· | ···A················ |
| O | 3650 | hCoV-19/England/ALDP-A6D9FD/2020\|EPI_ISL_607274\|20 | ···················· | ···················· | ····················· | ···················· | ···················· | ···················· |
| O | 3651 | hCoV-19/England/ALDP-A6EC6C/2020\|EPI_ISL_607275\|20 | ···················· | ···················· | ····················· | ···················· | ···················· | ···················· |
| O | 3652 | hCoV-19/England/ALDP-A6C7D6/2020\|EPI_ISL_607276\|20 | ···················· | ···················· | ····················· | ···················· | ···················· | ···················· |
| O | 3653 | hCoV-19/England/ALDP-A6EB41/2020\|EPI_ISL_607277\|20 | ···················· | ···················· | ····················· | ···················· | ···················· | ···················· |
| O | 3654 | hCoV-19/England/ALDP-A6F23C/2020\|EPI_ISL_607278\|20 | ···················· | ···················· | ····················· | ···················· | ···················· | ···················· |
| O | 3655 | hCoV-19/England/ALDP-A6F287/2020\|EPI_ISL_607279\|20 | ···················· | ···················· | ····················· | ···················· | ···················· | ···················· |
| O | 3656 | hCoV-19/England/ALDP-A6CD2F/2020\|EPI_ISL_607281\|20 | ···················· | ···················· | ····················· | ···················· | ···················· | ···················· |
| O | 3657 | hCoV-19/England/ALDP-A6F8FE/2020\|EPI_ISL_607282\|20 | ···················· | ···················· | ····················· | ···················· | ···················· | ···················· |
| O | 3658 | hCoV-19/England/ALDP-A6F2B4/2020\|EPI_ISL_607283\|20 | ···················· | ···················· | ····················· | ···················· | ···················· | ···················· |
| O | 3659 | hCoV-19/England/ALDP-A6F24B/2020\|EPI_ISL_607284\|20 | ···················· | ···················· | ····················· | ···················· | ···················· | ···················· |
| O | 3660 | hCoV-19/England/ALDP-A6F8EF/2020\|EPI_ISL_607285\|20 | ···················· | ···················· | ····················· | ···················· | ···················· | ···················· |
| O | 3661 | hCoV-19/England/ALDP-A6CC22/2020\|EPI_ISL_607286\|20 | ···················· | ···················· | ····················· | ···················· | ···················· | ···················· |
| O | 3662 | hCoV-19/England/ALDP-A6E58F/2020\|EPI_ISL_607287\|20 | ···················· | ···················· | ····················· | ···················· | ···················· | ···················· |
| O | 3663 | hCoV-19/England/ALDP-A6E701/2020\|EPI_ISL_607288\|20 | ···················· | ···················· | ····················· | ···················· | ···················· | ···················· |
| O | 3664 | hCoV-19/England/ALDP-A6EAF9/2020\|EPI_ISL_607289\|20 | ···················· | ···················· | ····················· | ···················· | ···················· | ···················· |
| O | 3665 | hCoV-19/England/ALDP-A6DCB8/2020\|EPI_ISL_607290\|20 | ···················· | ···················· | ····················· | ···················· | ···················· | ···················· |
| O | 3666 | hCoV-19/England/MILK-A7B26A/2020\|EPI_ISL_607292\|20 | ···················· | ···················· | ····················· | ···················· | ···················· | ···················· |
| O | 3667 | hCoV-19/England/MILK-A7BA53/2020\|EPI_ISL_607293\|20 | ···················· | ···················· | ····················· | ···················· | ···················· | ···················· |
| O | 3668 | hCoV-19/England/MILK-A7C7C4/2020\|EPI_ISL_607294\|20 | ···················· | ···················· | ····················· | ···················· | ···················· | ···················· |
| O | 3669 | hCoV-19/England/MILK-A7B73E/2020\|EPI_ISL_607295\|20 | ···················· | ···················· | ····················· | ···················· | ···················· | ···················· |
| O | 3670 | hCoV-19/England/MILK-A7B40A/2020\|EPI_ISL_607296\|20 | ···················· | ···················· | ····················· | ···················· | ···················· | ···················· |
| O | 3671 | hCoV-19/England/MILK-A7B024/2020\|EPI_ISL_607297\|20 | ···················· | ·A·················· | ····················· | ···················· | ···················· | ···A················ |
| O | 3672 | hCoV-19/England/MILK-A7B4A0/2020\|EPI_ISL_607298\|20 | ···················· | ···················· | ····················· | ···················· | ···················· | ···················· |
| O | 3673 | hCoV-19/England/MILK-A7B08E/2020\|EPI_ISL_607299\|20 | ···················· | ···················· | ····················· | ···················· | ···················· | ···················· |
| O | 3674 | hCoV-19/England/ALDP-A67F5B/2020\|EPI_ISL_608520\|20 | ···················· | ···················· | ····················· | ···················· | ···················· | ···················· |
| O | 3675 | hCoV-19/England/CAMC-A65A4D/2020\|EPI_ISL_608521\|20 | ···················· | ···················· | ····················· | ···················· | ···················· | ···················· |
| O | 3676 | hCoV-19/England/CAMC-A6500F/2020\|EPI_ISL_608522\|20 | ···················· | ···················· | ····················· | ···················· | ···················· | ···················· |
| O | 3677 | hCoV-19/England/CAMC-A658CB/2020\|EPI_ISL_608524\|20 | ···················· | ···················· | ····················· | ···················· | ···················· | ···················· |
| O | 3678 | hCoV-19/England/ALDP-A68FC3/2020\|EPI_ISL_608526\|20 | ···················· | ···················· | ····················· | ···················· | ···················· | ···················· |
| O | 3679 | hCoV-19/England/CAMC-A6501E/2020\|EPI_ISL_608527\|20 | ···················· | ···················· | ····················· | ···················· | ···················· | ···················· |
| O | 3680 | hCoV-19/England/ALDP-A68840/2020\|EPI_ISL_608528\|20 | ···················· | ···················· | ····················· | ···················· | ···················· | ···················· |
| O | 3681 | hCoV-19/England/CAMC-A6556A/2020\|EPI_ISL_608529\|20 | ···················· | ···················· | ····················· | ···················· | ···················· | ···················· |
| O | 3682 | hCoV-19/England/CAMC-A64B87/2020\|EPI_ISL_608530\|20 | ···················· | ···················· | ····················· | ···················· | ······K············· | ···················· |
| O | 3683 | hCoV-19/England/CAMC-A651DF/2020\|EPI_ISL_608531\|20 | ···················· | ···················· | ····················· | ···················· | ···················· | ···················· |
| O | 3684 | hCoV-19/England/CAMC-A64B1E/2020\|EPI_ISL_608532\|20 | ···················· | ·A·················· | ····················· | ···················· | ···················· | ···A················ |
| O | 3685 | hCoV-19/England/ALDP-A67EE5/2020\|EPI_ISL_608533\|20 | ···················· | ···················· | ····················· | ···················· | ···················· | ···················· |
| O | 3686 | hCoV-19/England/ALDP-A6878F/2020\|EPI_ISL_608534\|20 | ···················· | ···················· | ····················· | ···················· | ···················· | ···················· |
| O | 3687 | hCoV-19/England/CAMC-A64D90/2020\|EPI_ISL_608535\|20 | ···················· | ···················· | ····················· | ···················· | ···················· | ···················· |
| O | 3688 | hCoV-19/England/CAMC-A6562B/2020\|EPI_ISL_608536\|20 | ···················· | ···················· | ····················· | ···················· | ···················· | ···················· |
| O | 3689 | hCoV-19/England/CAMC-A65166/2020\|EPI_ISL_608537\|20 | ···················· | ···················· | ····················· | ···················· | ···················· | ···················· |
| O | 3690 | hCoV-19/England/CAMC-A655E2/2020\|EPI_ISL_608540\|20 | ···················· | ·A·················· | ····················· | ···················· | ···················· | ···················· |
| O | 3691 | hCoV-19/England/ALDP-A68EC6/2020\|EPI_ISL_608541\|20 | ···················· | ···················· | ····················· | ···················· | ···················· | ···················· |
| O | 3692 | hCoV-19/England/ALDP-A687F8/2020\|EPI_ISL_608542\|20 | ···················· | ···················· | ····················· | ···················· | ···················· | ···················· |
| O | 3693 | hCoV-19/England/CAMC-A6539D/2020\|EPI_ISL_608544\|20 | ···················· | ···················· | ····················· | ···················· | ···················· | ···················· |
| O | 3694 | hCoV-19/England/CAMC-A65A7A/2020\|EPI_ISL_608545\|20 | ···················· | ···················· | ····················· | ···················· | ···················· | ···················· |
| O | 3695 | hCoV-19/England/CAMC-A64BF0/2020\|EPI_ISL_608547\|20 | ···················· | ···················· | ····················· | ···················· | ···················· | ···················· |
| O | 3696 | hCoV-19/England/ALDP-A68822/2020\|EPI_ISL_608548\|20 | ···················· | ···················· | ····················· | ···················· | ···················· | ···················· |
| O | 3697 | hCoV-19/England/ALDP-A68145/2020\|EPI_ISL_608549\|20 | ···················· | ···················· | ····················· | ···················· | ···················· | ···················· |
| O | 3698 | hCoV-19/England/CAMC-A654E5/2020\|EPI_ISL_608550\|20 | ···················· | ···················· | ····················· | ···················· | ···················· | ···················· |
| O | 3699 | hCoV-19/England/ALDP-A6850D/2020\|EPI_ISL_608551\|20 | ···················· | ···················· | ····················· | ···················· | ···················· | ···················· |
| O | 3700 | hCoV-19/England/CAMC-A64D72/2020\|EPI_ISL_608552\|20 | ···················· | ···················· | ····················· | ···················· | ···················· | ···················· |
| O | 3701 | hCoV-19/England/ALDP-A6843D/2020\|EPI_ISL_608553\|20 | ···················· | ···················· | ····················· | ···················· | ···················· | ···················· |
| O | 3702 | hCoV-19/England/ALDP-A68CDB/2020\|EPI_ISL_608554\|20 | ···················· | ···················· | ····················· | ···················· | ···················· | ···················· |
| O | 3703 | hCoV-19/England/ALDP-A67F00/2020\|EPI_ISL_608556\|20 | ···················· | ···················· | ····················· | ···················· | ···················· | ···················· |
| O | 3704 | hCoV-19/England/CAMC-A64E33/2020\|EPI_ISL_608557\|20 | ···················· | ·A·················· | ····················· | ···················· | ···················· | ···················· |
| O | 3705 | hCoV-19/England/CAMC-A64D54/2020\|EPI_ISL_608558\|20 | ···················· | ·········A·········· | ····················· | ···················· | ···················· | ···················· |
| O | 3706 | hCoV-19/England/CAMC-A65193/2020\|EPI_ISL_608559\|20 | ···················· | ···················· | ····················· | ···················· | ···················· | ···················· |
| O | 3707 | hCoV-19/England/CAMC-A64C0C/2020\|EPI_ISL_608561\|20 | ···················· | ···················· | ····················· | ···················· | ······T············· | ···················· |
| O | 3708 | hCoV-19/England/ALDP-A67B66/2020\|EPI_ISL_608563\|20 | ···················· | ······C············· | ····················· | ···················· | ···················· | ···················· |
| O | 3709 | hCoV-19/England/ALDP-A67E21/2020\|EPI_ISL_608564\|20 | ···················· | ···················· | ····················· | ···················· | ···················· | ···················· |
| O | 3710 | hCoV-19/England/CAMC-A65139/2020\|EPI_ISL_608565\|20 | ···················· | ···················· | ····················· | ···················· | ···················· | ···················· |
| O | 3711 | hCoV-19/England/CAMC-A65B77/2020\|EPI_ISL_608566\|20 | ···················· | ···················· | ····················· | ···················· | ···················· | ···················· |
| O | 3712 | hCoV-19/England/ALDP-A68E5D/2020\|EPI_ISL_608567\|20 | ···················· | ···················· | ····················· | ···················· | ···················· | ···················· |
| O | 3713 | hCoV-19/England/CAMC-A656FE/2020\|EPI_ISL_608568\|20 | ···················· | ···················· | ····················· | ···················· | ···················· | ···················· |
| O | 3714 | hCoV-19/England/ALDP-A691BD/2020\|EPI_ISL_608569\|20 | ···················· | ···················· | ····················· | ···················· | ···················· | ···················· |
| O | 3715 | hCoV-19/England/CAMC-A65B68/2020\|EPI_ISL_608570\|20 | ···················· | ···················· | ····················· | ···················· | ···················· | ···················· |

O 3716 hCoV-19/England/CAMC-A6552E/2020|EPI_ISL_608571|20 ···················· ···················· ····················· ···················· ···················· ····················
O 3717 hCoV-19/England/CAMC-A654D6/2020|EPI_ISL_608572|20 ···················· ···················· ····················· ···················· ···················· ····················
O 3718 hCoV-19/England/ALDP-A6853A/2020|EPI_ISL_608575|20 ···················· ···················· ····················· ···················· ···················· ····················
O 3719 hCoV-19/England/CAMC-A6563A/2020|EPI_ISL_608576|20 ···················· ···················· ····················· ···················· ···················· ····················
O 3720 hCoV-19/England/CAMC-A64CFD/2020|EPI_ISL_608577|20 ···················· ···················· ····················· ···················· ···················· ····················
O 3721 hCoV-19/England/CAMC-A65B59/2020|EPI_ISL_608578|20 ···················· ···················· ····················· ···················· ···················· ····················
O 3722 hCoV-19/England/ALDP-A68813/2020|EPI_ISL_608579|20 ···················· ···················· ····················· ···················· ···················· ····················
O 3723 hCoV-19/England/ALDP-A688D7/2020|EPI_ISL_608580|20 ···················· ···················· ····················· ···················· ···················· ····················
O 3724 hCoV-19/England/ALDP-A68FB4/2020|EPI_ISL_608581|20 ···················· ···················· ····················· ···················· ···················· ····················
O 3725 hCoV-19/England/ALDP-A68FD2/2020|EPI_ISL_608582|20 ···················· ···················· ····················· ···················· ···················· ····················
O 3726 hCoV-19/England/ALDP-A68B38/2020|EPI_ISL_608583|20 ···················· ···················· ····················· ···················· ···················· ····················
O 3727 hCoV-19/England/ALDP-A681FA/2020|EPI_ISL_608584|20 ···················· ···················· ····················· ···················· ···················· ····················
O 3728 hCoV-19/England/ALDP-A681BE/2020|EPI_ISL_608586|20 ···················· ···················· ····················· ···················· ···················· ····················
O 3729 hCoV-19/England/ALDP-A68BFC/2020|EPI_ISL_608587|20 ···················· ···················· ····················· ···················· ···················· ····················
O 3730 hCoV-19/England/CAMC-A651FD/2020|EPI_ISL_608588|20 ···················· ···················· ····················· ···················· ···················· ····················
O 3731 hCoV-19/England/ALDP-A68BED/2020|EPI_ISL_608589|20 ···················· ···················· ····················· ···················· ···················· ····················
O 3732 hCoV-19/England/ALDP-A68497/2020|EPI_ISL_608590|20 ···················· ···················· ····················· ···················· ···················· ····················
O 3733 hCoV-19/England/ALDP-A67EB8/2020|EPI_ISL_608591|20 ···················· ···················· ····················· ···················· ···················· ····················
O 3734 hCoV-19/England/ALDP-A685D0/2020|EPI_ISL_608592|20 ···················· ···················· ····················· ···················· ···················· ····················
O 3735 hCoV-19/England/ALDP-A67ED6/2020|EPI_ISL_608593|20 ···················· ···················· ····················· ···················· ···················· ····················
O 3736 hCoV-19/England/ALDP-A68F2D/2020|EPI_ISL_608594|20 ···················· ···················· ····················· ···················· ···················· ····················
O 3737 hCoV-19/England/ALDP-A68488/2020|EPI_ISL_608595|20 ···················· ···················· ····················· ···················· ···················· ····················
O 3738 hCoV-19/England/CAMC-A64B0F/2020|EPI_ISL_608597|20 ···················· ···················· ····················· ···················· ···················· ····················
O 3739 hCoV-19/England/CAMC-A656A3/2020|EPI_ISL_608598|20 ···················· ···················· ····················· ···················· ···················· ····················
O 3740 hCoV-19/England/ALDP-A68F3C/2020|EPI_ISL_608599|20 ···················· ···················· ····················· ···················· ···················· ····················
O 3741 hCoV-19/England/204121801/2020|EPI_ISL_609850|2020 ···················· ···················· ····················· ···················· ···················· ····················
O 3742 hCoV-19/England/204121805/2020|EPI_ISL_609851|2020 ···················· ···················· ····················· ···················· ···················· ····················
O 3743 hCoV-19/England/204141598/2020|EPI_ISL_609852|2020 ···················· ·A·················· ····················· ···················· ···················· ··A·················
O 3744 hCoV-19/England/204161206/2020|EPI_ISL_609853|2020 ···················· ···················· ····················· ···················· ···················· ····················
O 3745 hCoV-19/England/204161234/2020|EPI_ISL_609854|2020 ···················· ···················· ····················· ···················· ···················· ····················
O 3746 hCoV-19/England/204180580/2020|EPI_ISL_609855|2020 ···················· ···················· ····················· ···················· ···················· ····················
O 3747 hCoV-19/England/204180611/2020|EPI_ISL_609856|2020 ···················· ···················· ····················· ···················· ···················· ····················
O 3748 hCoV-19/England/204180614/2020|EPI_ISL_609857|2020 ···················· ················T··· ····················· ···················· ···················· ····················
O 3749 hCoV-19/England/204190389/2020|EPI_ISL_609858|2020 ···················· ···················· ····················· ···················· ···················· ····················
O 3750 hCoV-19/England/204191235/2020|EPI_ISL_609859|2020 ···················· ···················· ····················· ···················· ···················· ····················
O 3751 hCoV-19/England/204191267/2020|EPI_ISL_609860|2020 ···················· ···················· ····················· ···················· ···················· ····················
O 3752 hCoV-19/England/204191532/2020|EPI_ISL_609861|2020 ···················· ·A·················· ····················· ···················· ···················· ··A·················
O 3753 hCoV-19/England/204200094/2020|EPI_ISL_609863|2020 ···················· ···················· ····················· ···················· ···················· ····················
O 3754 hCoV-19/England/204200095/2020|EPI_ISL_609864|2020 ···················· ···················· ····················· ···················· ···················· ····················
O 3755 hCoV-19/England/204200096/2020|EPI_ISL_609865|2020 ···················· ···················· ····················· ···················· ···················· ····················
O 3756 hCoV-19/England/204200098/2020|EPI_ISL_609866|2020 ···················· ···················· ····················· ···················· ···················· ····················
O 3757 hCoV-19/England/204200106/2020|EPI_ISL_609867|2020 ···················· ···················· ····················· ···················· ···················· ····················
O 3758 hCoV-19/England/204200112/2020|EPI_ISL_609868|2020 ···················· ···················· ····················· ···················· ···················· ····················
O 3759 hCoV-19/England/204200157/2020|EPI_ISL_609869|2020 ···················· ···················· ····················· ···················· ···················· ····················
O 3760 hCoV-19/England/204200161/2020|EPI_ISL_609870|2020 ···················· ···················· ····················· ···················· ···················· ····················
O 3761 hCoV-19/England/204200163/2020|EPI_ISL_609871|2020 ···················· ···················· ····················· ···················· ···················· ····················
O 3762 hCoV-19/England/204200172/2020|EPI_ISL_609875|2020 ···················· ···················· ····················· ···················· ···················· ····················
O 3763 hCoV-19/England/204200187/2020|EPI_ISL_609877|2020 ···················· ···················· ····················· ···················· ···················· ····················
O 3764 hCoV-19/England/204200188/2020|EPI_ISL_609878|2020 ···················· ···················· ····················· ···················· ·A·················· ····················
O 3765 hCoV-19/England/204200191/2020|EPI_ISL_609879|2020 ···················· ···················· ····················· ···················· ···················· ····················
O 3766 hCoV-19/England/204200194/2020|EPI_ISL_609880|2020 ···················· ···················· ····················· ···················· ···················· ····················
O 3767 hCoV-19/England/204200416/2020|EPI_ISL_609881|2020 ···················· ···················· ····················· ···················· ···················· ····················
O 3768 hCoV-19/England/204200491/2020|EPI_ISL_609882|2020 ···················· ···················· ····················· ···················· ···················· ····················
O 3769 hCoV-19/England/204200592/2020|EPI_ISL_609883|2020 ···················· ···················· ····················· ···················· ···················· ····················
O 3770 hCoV-19/England/204200600/2020|EPI_ISL_609884|2020 ···················· ···················· ····················· ···················· ···················· ····················
O 3771 hCoV-19/England/204220384/2020|EPI_ISL_609885|2020 ···················· ···················· ····················· ···················· ···················· ····················
O 3772 hCoV-19/England/204220385/2020|EPI_ISL_609886|2020 ···················· ···················· ····················· ···················· ···················· ····················
O 3773 hCoV-19/England/204220386/2020|EPI_ISL_609887|2020 ···················· ···················· ····················· ···················· ···················· ····················
O 3774 hCoV-19/England/204220387/2020|EPI_ISL_609888|2020 ···················· ···················· ····················· ···················· ···················· ····················
O 3775 hCoV-19/England/204220905/2020|EPI_ISL_609889|2020 ···················· ···················· ····················· ···················· ···················· ····················
O 3776 hCoV-19/England/204240440/2020|EPI_ISL_609892|2020 ···················· ···················· ····················· ···················· ···················· ····················
O 3777 hCoV-19/England/204241000/2020|EPI_ISL_609894|2020 ···················· ···················· ····················· ···················· ···················· ····················
O 3778 hCoV-19/England/204241008/2020|EPI_ISL_609895|2020 ···················· ···················· ····················· ···················· ···················· ····················
O 3779 hCoV-19/England/204241197/2020|EPI_ISL_609896|2020 ···················· ···················· ····················· ···················· ···················· ····················
O 3780 hCoV-19/England/204241198/2020|EPI_ISL_609897|2020 ········Y··········· ···················· ····················· ···················· ···················· ····················
O 3781 hCoV-19/England/204260115/2020|EPI_ISL_609898|2020 ···················· ···················· ····················· ···················· ···················· ····················
O 3782 hCoV-19/England/204260213/2020|EPI_ISL_609899|2020 ···················· ···················· ····················· ···················· ···················· ····················
O 3783 hCoV-19/England/CAMC-A81726/2020|EPI_ISL_610832|20 ···················· ·A·················· ····················· ···················· ···················· ····················
O 3784 hCoV-19/England/CAMC-A81B2A/2020|EPI_ISL_610833|20 ···················· ···················· ····················· ···················· ···················· ····················
O 3785 hCoV-19/England/CAMC-A80D9E/2020|EPI_ISL_610835|20 ···················· ···················· ····················· ···················· ···················· ····················
O 3786 hCoV-19/England/CAMC-A82A0E/2020|EPI_ISL_610836|20 ···················· ···················· ····················· ···················· ···················· ····················
O 3787 hCoV-19/England/CAMC-A81DCA/2020|EPI_ISL_610837|20 ···················· ···················· ····················· ···················· ···················· ····················
O 3788 hCoV-19/England/CAMC-A8300B/2020|EPI_ISL_610838|20 ···················· ···················· ····················· ···················· ······T············· ····················
O 3789 hCoV-19/England/CAMC-A82998/2020|EPI_ISL_610839|20 ···················· ···················· ····················· ···················· ···················· ····················
O 3790 hCoV-19/England/CAMC-A81FE2/2020|EPI_ISL_610840|20 ···················· ···················· ····················· ···················· ···················· ····················
O 3791 hCoV-19/England/CAMC-A811EC/2020|EPI_ISL_610841|20 ···················· ···················· ····················· ···················· ······T············· ····················
O 3792 hCoV-19/England/CAMC-A81F3D/2020|EPI_ISL_610842|20 ···················· ···················· ····················· ···················· ···················· ····················
O 3793 hCoV-19/England/CAMC-A83029/2020|EPI_ISL_610844|20 ···················· ···················· ····················· ···················· ······T············· ····················
O 3794 hCoV-19/England/CAMC-A816FC/2020|EPI_ISL_610845|20 ···················· ···················· ····················· ···················· ······T············· ····················
O 3795 hCoV-19/England/CAMC-A82E8A/2020|EPI_ISL_610846|20 ···················· ···················· ····················· ···················· ···················· ····················

| | | | | | | | |
|---|---|---|---|---|---|---|---|
| O | 3796 hCoV-19/England/CAMC-A82251/2020\|EPI_ISL_610847\|20 | ···················· | ···················· | ····················· | ···················· | ···················· | ···················· |
| O | 3797 hCoV-19/England/CAMC-A80D16/2020\|EPI_ISL_610848\|20 | ···················· | ···················· | ····················· | ···················· | ···················· | ···················· |
| O | 3798 hCoV-19/England/CAMC-A82FE1/2020\|EPI_ISL_610849\|20 | ···················· | ···················· | ····················· | ···················· | ···················· | ···················· |
| O | 3799 hCoV-19/England/CAMC-A810FE/2020\|EPI_ISL_610850\|20 | ···················· | ···················· | ····················· | ···················· | ···················· | ···················· |
| O | 3800 hCoV-19/England/CAMC-A821FA/2020\|EPI_ISL_610852\|20 | ···················· | ·A·················· | ····················· | ···················· | ···················· | ··A················· |
| O | 3801 hCoV-19/England/CAMC-A82F5A/2020\|EPI_ISL_610854\|20 | ···················· | ···················· | ····················· | ···················· | ···················· | ···················· |
| O | 3802 hCoV-19/England/CAMC-A80CEC/2020\|EPI_ISL_610855\|20 | ···················· | ···················· | ····················· | ···················· | ···················· | ···················· |
| O | 3803 hCoV-19/England/CAMC-A8239A/2020\|EPI_ISL_610856\|20 | ···················· | ···················· | ····················· | ···················· | ···················· | ···················· |
| O | 3804 hCoV-19/England/CAMC-A8293E/2020\|EPI_ISL_610857\|20 | ···················· | ···················· | ····················· | ···················· | ···················· | ···················· |
| O | 3805 hCoV-19/England/CAMC-A82181/2020\|EPI_ISL_610859\|20 | ···················· | ···················· | ····················· | ···················· | ···················· | ···················· |
| O | 3806 hCoV-19/England/CAMC-A82FA5/2020\|EPI_ISL_610860\|20 | ···················· | ···················· | ····················· | ···················· | ···················· | ···················· |
| O | 3807 hCoV-19/England/CAMC-A810B2/2020\|EPI_ISL_610862\|20 | ···················· | ···················· | ····················· | ···················· | ···················· | ···················· |
| O | 3808 hCoV-19/England/CAMC-A81182/2020\|EPI_ISL_610863\|20 | ···················· | ···················· | ····················· | ···················· | ···················· | ···················· |
| O | 3809 hCoV-19/England/CAMC-A819B7/2020\|EPI_ISL_610864\|20 | ···················· | ···················· | ····················· | ···················· | ···················· | ···················· |
| O | 3810 hCoV-19/England/CAMC-A80D07/2020\|EPI_ISL_610865\|20 | ···················· | ···················· | ····················· | ···················· | ···················· | ···················· |
| O | 3811 hCoV-19/England/CAMC-A811CE/2020\|EPI_ISL_610866\|20 | ···················· | ···················· | ····················· | ···················· | ···················· | ···················· |
| O | 3812 hCoV-19/England/CAMC-A82A86/2020\|EPI_ISL_610867\|20 | ···················· | ···················· | ····················· | ···················· | ···················· | ···················· |
| O | 3813 hCoV-19/England/CAMC-A82F2D/2020\|EPI_ISL_610869\|20 | ···················· | ···················· | ····················· | ···················· | ···················· | ···················· |
| O | 3814 hCoV-19/England/CAMC-A816DE/2020\|EPI_ISL_610870\|20 | ···················· | ···················· | ····················· | ···················· | ···················· | ···················· |
| O | 3815 hCoV-19/England/CAMC-A82F69/2020\|EPI_ISL_610871\|20 | ···················· | ···················· | ····················· | ···················· | ···················· | ···················· |
| O | 3816 hCoV-19/England/CAMC-A82039/2020\|EPI_ISL_610872\|20 | ···················· | ···················· | ····················· | ···················· | ···················· | ···················· |
| O | 3817 hCoV-19/England/CAMC-A83074/2020\|EPI_ISL_610873\|20 | ···················· | ·A·················· | ····················· | ···················· | ···················· | ··A················· |
| O | 3818 hCoV-19/England/CAMC-A819D5/2020\|EPI_ISL_610874\|20 | ···················· | ···················· | ····················· | ···················· | ···················· | ···················· |
| O | 3819 hCoV-19/England/CAMC-A823E5/2020\|EPI_ISL_610875\|20 | ···················· | ···················· | ····················· | ···················· | ···················· | ···················· |
| O | 3820 hCoV-19/England/CAMC-A82E99/2020\|EPI_ISL_610877\|20 | ···················· | ···················· | ····················· | ···················· | ···················· | ···················· |
| O | 3821 hCoV-19/England/CAMC-A81C27/2020\|EPI_ISL_610878\|20 | ···················· | ···················· | ····················· | ···················· | ···················· | ···················· |
| O | 3822 hCoV-19/England/CAMC-A8296B/2020\|EPI_ISL_610879\|20 | ···················· | ···················· | ····················· | ···················· | ···················· | ···················· |
| O | 3823 hCoV-19/England/CAMC-A81146/2020\|EPI_ISL_610881\|20 | ···················· | ···················· | ····················· | ···················· | ···················· | ···················· |
| O | 3824 hCoV-19/England/CAMC-A81EC7/2020\|EPI_ISL_610883\|20 | ···················· | ···················· | ····················· | ···················· | ······T············· | ···················· |
| O | 3825 hCoV-19/England/CAMC-A82734/2020\|EPI_ISL_610884\|20 | ···················· | ···················· | ····················· | ···················· | ···················· | ···················· |
| O | 3826 hCoV-19/England/CAMC-A80CDD/2020\|EPI_ISL_610886\|20 | ···················· | ···················· | ····················· | ···················· | ···················· | ···················· |
| O | 3827 hCoV-19/England/CAMC-A81E4F/2020\|EPI_ISL_610887\|20 | ···················· | ···················· | ····················· | ···················· | ···················· | ···················· |
| O | 3828 hCoV-19/England/CAMC-A81E30/2020\|EPI_ISL_610888\|20 | ···················· | ···················· | ····················· | ···················· | ···················· | ···················· |
| O | 3829 hCoV-19/England/CAMC-A81F00/2020\|EPI_ISL_610889\|20 | ···················· | ···················· | ····················· | ···················· | ···················· | ···················· |
| O | 3830 hCoV-19/England/CAMC-A822AC/2020\|EPI_ISL_610890\|20 | ···················· | ···················· | ····················· | ···················· | ···················· | ···················· |
| O | 3831 hCoV-19/England/CAMC-A80F7A/2020\|EPI_ISL_610891\|20 | ···················· | ···················· | ····················· | ···················· | ···················· | ···················· |
| O | 3832 hCoV-19/England/CAMC-A81173/2020\|EPI_ISL_610893\|20 | ···················· | ···················· | ····················· | ···················· | ···················· | ···················· |
| O | 3833 hCoV-19/England/CAMC-A81BB1/2020\|EPI_ISL_610894\|20 | ···················· | ···················· | ····················· | ···················· | ···················· | ···················· |
| O | 3834 hCoV-19/England/CAMC-A82215/2020\|EPI_ISL_610895\|20 | ···················· | ···················· | ····················· | ···················· | ···················· | ···················· |
| O | 3835 hCoV-19/England/CAMC-A81E21/2020\|EPI_ISL_610896\|20 | ···················· | ···················· | ····················· | ···················· | ···················· | ···················· |
| O | 3836 hCoV-19/England/CAMC-A81A96/2020\|EPI_ISL_610897\|20 | ···················· | ···················· | ····················· | ···················· | ···················· | ···················· |
| O | 3837 hCoV-19/England/CAMC-A8202A/2020\|EPI_ISL_610898\|20 | ···················· | ···················· | ····················· | ···················· | ···················· | ···················· |
| O | 3838 hCoV-19/Wales/PHWC-480DC1/2020\|EPI_ISL_613002\|2020 | ···················· | ···················· | ····················· | ···················· | ···················· | ···················· |
| O | 3839 hCoV-19/Wales/PHWC-480DEF/2020\|EPI_ISL_613004\|2020 | ···················· | ···················· | ····················· | ···················· | ···················· | ···················· |
| O | 3840 hCoV-19/Wales/PHWC-480DFE/2020\|EPI_ISL_613005\|2020 | ···················· | ···················· | ····················· | ···················· | ···················· | ···················· |
| O | 3841 hCoV-19/England/ALDP-A6E297/2020\|EPI_ISL_607110\|20 | ···················· | ···················· | ····················· | ···················· | ···················· | ···················· |
| O | 3842 hCoV-19/England/ALDP-A6F1C6/2020\|EPI_ISL_607111\|20 | ···················· | ···················· | ····················· | ···················· | ···················· | ···················· |
| O | 3843 hCoV-19/England/ALDP-A6EB05/2020\|EPI_ISL_607112\|20 | ···················· | ···················· | ····················· | ···················· | ···················· | ···················· |
| O | 3844 hCoV-19/England/ALDP-A6FC5C/2020\|EPI_ISL_607113\|20 | ···················· | ···················· | ····················· | ···················· | ···················· | ···················· |
| O | 3845 hCoV-19/England/ALDP-A6F15D/2020\|EPI_ISL_607114\|20 | ···················· | ···················· | ····················· | ···················· | ···················· | ···················· |
| O | 3846 hCoV-19/England/ALDP-A6C976/2020\|EPI_ISL_607115\|20 | ···················· | ···················· | ····················· | ···················· | ···················· | ···················· |
| O | 3847 hCoV-19/England/ALDP-A6F74C/2020\|EPI_ISL_607117\|20 | ···················· | ···················· | ····················· | ···················· | ···················· | ···················· |
| O | 3848 hCoV-19/England/ALDP-A6C7A9/2020\|EPI_ISL_607118\|20 | ···················· | ···················· | ····················· | ···················· | ···················· | ···················· |
| O | 3849 hCoV-19/England/ALDP-A6D948/2020\|EPI_ISL_607119\|20 | ···················· | ···················· | ····················· | ···················· | ···················· | ···················· |
| O | 3850 hCoV-19/England/ALDP-A6E6F5/2020\|EPI_ISL_607120\|20 | ···················· | ···················· | ····················· | ···················· | ···················· | ···················· |
| O | 3851 hCoV-19/England/ALDP-A6EB32/2020\|EPI_ISL_607121\|20 | ···················· | ···················· | ····················· | ···················· | ···················· | ···················· |
| O | 3852 hCoV-19/England/ALDP-A6CC8C/2020\|EPI_ISL_607122\|20 | ···················· | ···················· | ···········T········· | ···················· | ···················· | ···················· |
| O | 3853 hCoV-19/England/ALDP-A6EB7E/2020\|EPI_ISL_607123\|20 | ···················· | ···················· | ····················· | ···················· | ···················· | ···················· |
| O | 3854 hCoV-19/England/ALDP-A6F7D3/2020\|EPI_ISL_607124\|20 | ···················· | ···················· | ····················· | ···················· | ···················· | ···················· |
| O | 3855 hCoV-19/England/ALDP-A6F973/2020\|EPI_ISL_607125\|20 | ···················· | ···················· | ····················· | ···················· | ···················· | ···················· |
| O | 3856 hCoV-19/England/ALDP-A6EEC0/2020\|EPI_ISL_607126\|20 | ···················· | ···················· | ····················· | ···················· | ···················· | ···················· |
| O | 3857 hCoV-19/England/ALDP-A6E570/2020\|EPI_ISL_607127\|20 | ···················· | ···················· | ····················· | ···················· | ···················· | ···················· |
| O | 3858 hCoV-19/England/ALDP-A6CAFB/2020\|EPI_ISL_607128\|20 | ···················· | ···················· | ····················· | ···················· | ···················· | ···················· |
| O | 3859 hCoV-19/England/ALDP-A6D5EA/2020\|EPI_ISL_607129\|20 | ···················· | ···················· | ····················· | ···················· | ···················· | ···················· |
| O | 3860 hCoV-19/England/ALDP-A6F269/2020\|EPI_ISL_607130\|20 | ···················· | ···················· | ····················· | ···················· | ···················· | ···················· |
| O | 3861 hCoV-19/England/ALDP-A68910/2020\|EPI_ISL_608440\|20 | ···················· | ···················· | ····················· | ···················· | ···················· | ···················· |
| O | 3862 hCoV-19/England/ALDP-A6842E/2020\|EPI_ISL_608441\|20 | ···················· | ···················· | ····················· | ···················· | ···················· | ···················· |
| O | 3863 hCoV-19/England/CAMC-A658F8/2020\|EPI_ISL_608442\|20 | ···················· | ···················· | ····················· | ···················· | ···················· | ···················· |
| O | 3864 hCoV-19/England/ALDP-A68E3F/2020\|EPI_ISL_608443\|20 | ···················· | ···················· | ····················· | ···················· | ···················· | ···················· |
| O | 3865 hCoV-19/England/ALDP-A67BDF/2020\|EPI_ISL_608444\|20 | ···················· | ···················· | ····················· | ···················· | ···················· | ···················· |
| O | 3866 hCoV-19/England/ALDP-A69153/2020\|EPI_ISL_608445\|20 | ···················· | ···················· | ····················· | ···················· | ···················· | ···················· |
| O | 3867 hCoV-19/England/CAMC-A648AE/2020\|EPI_ISL_608446\|20 | ···················· | ···················· | ····················· | ···················· | ···················· | ···················· |
| O | 3868 hCoV-19/England/CAMC-A64FE5/2020\|EPI_ISL_608447\|20 | ···················· | ···················· | ····················· | ···················· | ···················· | ···················· |
| O | 3869 hCoV-19/England/ALDP-A683D6/2020\|EPI_ISL_608448\|20 | ···················· | ···················· | ····················· | ···················· | ···················· | ···················· |
| O | 3870 hCoV-19/England/CAMC-A64C39/2020\|EPI_ISL_608449\|20 | ···················· | ···················· | ····················· | ···················· | ···················· | ···················· |
| O | 3871 hCoV-19/England/CAMC-A64E8E/2020\|EPI_ISL_608450\|20 | ···················· | ···················· | ····················· | ···················· | ···················· | ···················· |
| O | 3872 hCoV-19/England/CAMC-A64EF7/2020\|EPI_ISL_608451\|20 | ···················· | ···················· | ····················· | ···················· | ···················· | ···················· |
| O | 3873 hCoV-19/England/ALDP-A68558/2020\|EPI_ISL_608452\|20 | ···················· | ···················· | ····················· | ···················· | ···················· | ···················· |
| O | 3874 hCoV-19/England/ALDP-A68585/2020\|EPI_ISL_608453\|20 | ···················· | ···················· | ····················· | ···················· | ···················· | ···················· |
| O | 3875 hCoV-19/England/ALDP-A68242/2020\|EPI_ISL_608454\|20 | ···················· | ···················· | ····················· | ···················· | ···················· | ···················· |

O 3876 hCoV-19/England/ALDP-A685A3/2020|EPI_ISL_608457|20 ···················· ···················· ···················· ···················· ···················· ····················
O 3877 hCoV-19/England/CAMC-A65430/2020|EPI_ISL_608458|20 ···················· ···················· ···················· ···················· ···················· ····················
O 3878 hCoV-19/England/CAMC-A651C0/2020|EPI_ISL_608459|20 ···················· ···················· ···················· ···················· ···················· ····················
O 3879 hCoV-19/England/ALDP-A68901/2020|EPI_ISL_608461|20 ···················· ···················· ···················· ···················· ···················· ····················
O 3880 hCoV-19/England/ALDP-A6852B/2020|EPI_ISL_608500|20 ···················· ···················· ···················· ···················· ···················· ····················
O 3881 hCoV-19/England/CAMC-A65A89/2020|EPI_ISL_608501|20 ···················· ···················· ···················· ···················· ···················· ····················
O 3882 hCoV-19/England/ALDP-A68224/2020|EPI_ISL_608502|20 ···················· ···················· ···················· ···················· ···················· ····················
O 3883 hCoV-19/England/ALDP-A6879E/2020|EPI_ISL_608503|20 ···················· ···················· ···················· ···················· ···················· ····················
O 3884 hCoV-19/England/ALDP-A691CC/2020|EPI_ISL_608505|20 ···················· ···················· ···················· ···················· ···················· ····················
O 3885 hCoV-19/England/CAMC-A6537F/2020|EPI_ISL_608506|20 ···················· ···················· ···················· ···················· ···················· ····················
O 3886 hCoV-19/England/CAMC-A64C1B/2020|EPI_ISL_608507|20 ···················· ···················· ···················· ···················· ······T············· ····················
O 3887 hCoV-19/England/CAMC-A64D63/2020|EPI_ISL_608508|20 ···················· ···················· ···················· ···················· ···················· ····················
O 3888 hCoV-19/England/CAMC-A656B2/2020|EPI_ISL_608509|20 ···················· ···················· ···················· ···················· ···················· ····················
O 3889 hCoV-19/England/CAMC-A6599B/2020|EPI_ISL_608510|20 ···················· ···················· ···················· ···················· ······T············· ····················
O 3890 hCoV-19/England/ALDP-A681DC/2020|EPI_ISL_608511|20 ···················· ···················· ···················· ···················· ···················· ····················
O 3891 hCoV-19/England/ALDP-A69162/2020|EPI_ISL_608512|20 ···················· ···················· ···················· ···················· ···················· ····················
O 3892 hCoV-19/England/ALDP-A68C44/2020|EPI_ISL_608513|20 ···················· ···················· ···················· ···················· ···················· ····················
O 3893 hCoV-19/England/CAMC-A64FF4/2020|EPI_ISL_608514|20 ···················· ·A·················· ···················· ···················· ···················· ···A················
O 3894 hCoV-19/England/CAMC-A6503C/2020|EPI_ISL_608515|20 ···················· ···················· ···················· ···················· ···················· ····················
O 3895 hCoV-19/England/ALDP-A68163/2020|EPI_ISL_608517|20 ···················· ···················· ···················· ···················· ···················· ····················
O 3896 hCoV-19/England/ALDP-A688F5/2020|EPI_ISL_608518|20 ···················· ···················· ···················· ···················· ···················· ····················
O 3897 hCoV-19/England/ALDP-A6895C/2020|EPI_ISL_608519|20 ···················· ···················· ···················· ···················· ······T············· ····················
O 3898 hCoV-19/England/CAMC-A3E3CA/2020|EPI_ISL_609770|20 ···················· ···················· ···················· ···················· ···················· ····················
O 3899 hCoV-19/England/CAMC-A3E412/2020|EPI_ISL_609771|20 ···················· ···················· ···················· ···················· ···················· ····················
O 3900 hCoV-19/England/CAMC-A3F55A/2020|EPI_ISL_609772|20 ···················· ···················· ···················· ···················· ···················· ····················
O 3901 hCoV-19/England/CAMC-A3E5A6/2020|EPI_ISL_609773|20 ···················· ···················· ···················· ···················· ···················· ····················
O 3902 hCoV-19/England/CAMC-A3E48B/2020|EPI_ISL_609774|20 ···················· ···················· ···················· ···················· ···················· ····················
O 3903 hCoV-19/England/CAMC-A3F6B1/2020|EPI_ISL_609775|20 ···················· ···················· ···················· ···················· ···················· ····················
O 3904 hCoV-19/England/CAMC-A3FB0D/2020|EPI_ISL_609776|20 ···················· ···················· ···················· ···················· ···················· ····················
O 3905 hCoV-19/England/CAMC-A3EEBA/2020|EPI_ISL_609777|20 ···················· ···················· ···················· ···················· ···················· ····················
O 3906 hCoV-19/England/CAMC-A3EB2C/2020|EPI_ISL_609779|20 ···················· ···················· ···················· ···················· ···················· ····················
O 3907 hCoV-19/England/CAMC-A3EFD5/2020|EPI_ISL_609780|20 ···················· ···················· ···················· ···················· ···················· ····················
O 3908 hCoV-19/England/CAMC-A3EBB3/2020|EPI_ISL_609781|20 ···················· ···················· ···················· ···················· ···················· ····················
O 3909 hCoV-19/England/CAMC-A3E45E/2020|EPI_ISL_609782|20 ···················· ···················· ···················· ···················· ···················· ····················
O 3910 hCoV-19/England/CAMC-A3E825/2020|EPI_ISL_609783|20 ···················· ···················· ···················· ···················· ···················· ····················
O 3911 hCoV-19/England/CAMC-A3F693/2020|EPI_ISL_609784|20 ···················· ···················· ···················· ···················· ···················· ····················
O 3912 hCoV-19/England/CAMC-A3F8BB/2020|EPI_ISL_609785|20 ···················· ···················· ···················· ···················· ···················· ····················
O 3913 hCoV-19/England/CAMC-A3FA79/2020|EPI_ISL_609786|20 ···················· ···················· ···················· ···················· ···················· ····················
O 3914 hCoV-19/England/CAMC-A3EC29/2020|EPI_ISL_609787|20 ···················· ···················· ···················· ···················· ···················· ····················
O 3915 hCoV-19/England/CAMC-A3EAB6/2020|EPI_ISL_609788|20 ···················· ···················· ···················· ···················· ···················· ····················
O 3916 hCoV-19/England/CAMC-A3F833/2020|EPI_ISL_609789|20 ···················· ···················· ···················· ···················· ···················· ····················
O 3917 hCoV-19/England/CAMC-A3E5E2/2020|EPI_ISL_609790|20 ···················· ···················· ···················· ···················· ···················· ····················
O 3918 hCoV-19/England/CAMC-A3F8F7/2020|EPI_ISL_609791|20 ···················· ···················· ···················· ···················· ······T············· ····················
O 3919 hCoV-19/England/CAMC-A3F99A/2020|EPI_ISL_609792|20 ···················· ···················· ···················· ···················· ···················· ····················
O 3920 hCoV-19/England/CAMC-A3EA6B/2020|EPI_ISL_609800|20 ···················· ···················· ···················· ···················· ···················· ····················
O 3921 hCoV-19/Canada/ON-UHTC_0335/2020|EPI_ISL_609802|20 ···················· ···················· ···················· ···················· ···················· ····················
O 3922 hCoV-19/Canada/ON-UHTC_0345/2020|EPI_ISL_609804|20 ···················· ···················· ···················· ···················· ···················· ····················
O 3923 hCoV-19/Canada/ON-UHTC_0327/2020|EPI_ISL_609806|20 ···················· ·A·················· ···················· ···················· ···················· ···A················
O 3924 hCoV-19/Canada/ON-UHTC_0330/2020|EPI_ISL_609807|20 ···················· ···················· ···················· ···················· ···················· ····················
O 3925 hCoV-19/Canada/ON-UHTC_0341/2020|EPI_ISL_609808|20 ···················· ···················· ···················· ···················· ···················· ····················
O 3926 hCoV-19/Canada/ON-UHTC_0328/2020|EPI_ISL_609809|20 ···················· ···················· ···················· ···················· ···················· ····················
O 3927 hCoV-19/Canada/ON-UHTC_0356/2020|EPI_ISL_609811|20 ···················· ·A·················· ···················· ···················· ···················· ···A················
O 3928 hCoV-19/Canada/ON-UHTC_0331/2020|EPI_ISL_609813|20 ···················· ···················· ···················· ···················· ···················· ····················
O 3929 hCoV-19/Canada/ON-UHTC_0346/2020|EPI_ISL_609819|20 ···················· ···················· ···················· ···················· ···················· ····················
O 3930 hCoV-19/Canada/ON-UHTC_0324/2020|EPI_ISL_609820|20 ···················· ···················· ···················· ···················· ···················· ····················
O 3931 hCoV-19/Canada/ON-UHTC_0318/2020|EPI_ISL_609821|20 ···················· ···················· ···················· ···················· ···················· ····················
O 3932 hCoV-19/Canada/ON-UHTC_0355/2020|EPI_ISL_609822|20 ···················· ···················· ···················· ···················· ···················· ····················
O 3933 hCoV-19/Canada/ON-UHTC_0322/2020|EPI_ISL_609826|20 ···················· ·A·················· ···················· ···················· ···················· ···A················
O 3934 hCoV-19/England/204080122/2020|EPI_ISL_609827|2020 ···················· ···················· ···················· ···················· ···················· ····················
O 3935 hCoV-19/England/204080876/2020|EPI_ISL_609828|2020 ···················· ···················· ···················· ···················· ···················· ····················
O 3936 hCoV-19/England/204100232/2020|EPI_ISL_609833|2020 ···················· ···················· ···················· ···················· ···················· ····················
O 3937 hCoV-19/England/204100238/2020|EPI_ISL_609834|2020 ········Y··········· ···················· ···················· ···················· ···················· ····················
O 3938 hCoV-19/England/204100511/2020|EPI_ISL_609835|2020 ···················· ···················· ···················· ···················· ···················· ····················
O 3939 hCoV-19/England/204100512/2020|EPI_ISL_609836|2020 ···················· ···················· ···················· ···················· ···················· ····················
O 3940 hCoV-19/England/204100517/2020|EPI_ISL_609838|2020 ···················· ···················· ···················· ···················· ···················· ····················
O 3941 hCoV-19/England/204100524/2020|EPI_ISL_609839|2020 ···················· ···················· ···················· ···················· ···················· ····················
O 3942 hCoV-19/England/204100966/2020|EPI_ISL_609840|2020 ···················· ·A·················· ···················· ···················· ···················· ···A················
O 3943 hCoV-19/England/204100971/2020|EPI_ISL_609841|2020 ···················· ·A·················· ···················· ···················· ···················· ···A················
O 3944 hCoV-19/England/204100982/2020|EPI_ISL_609842|2020 ···················· ·A·················· ···················· ···················· ···················· ···A················
O 3945 hCoV-19/England/204120186/2020|EPI_ISL_609843|2020 ···················· ·A·················· ···················· ···················· ···················· ···A················
O 3946 hCoV-19/England/204120548/2020|EPI_ISL_609844|2020 ···················· ·A·················· ···················· ···················· ···················· ····················
O 3947 hCoV-19/England/204121763/2020|EPI_ISL_609845|2020 ···················· ···················· ···················· ···················· ···················· ····················
O 3948 hCoV-19/England/204121765/2020|EPI_ISL_609846|2020 ···················· ···················· ···················· ···················· ···················· ····················
O 3949 hCoV-19/England/204121769/2020|EPI_ISL_609847|2020 ···················· ···················· ···················· ···················· ···················· ····················
O 3950 hCoV-19/England/204121781/2020|EPI_ISL_609848|2020 ···················· ···················· ···················· ···················· ···················· ····················
O 3951 hCoV-19/England/204121782/2020|EPI_ISL_609849|2020 ···················· ···················· ···················· ···················· ···················· ····················
O 3952 hCoV-19/England/CAMC-A81C90/2020|EPI_ISL_610755|20 ···················· ···················· ···················· ···················· ···················· ····················
O 3953 hCoV-19/England/CAMC-A81BDF/2020|EPI_ISL_610756|20 ···················· ···················· ···················· ···················· ···················· ····················
O 3954 hCoV-19/England/CAMC-A8200C/2020|EPI_ISL_610757|20 ···················· ···················· ···················· ···················· ···················· ····················
O 3955 hCoV-19/England/CAMC-A80CCE/2020|EPI_ISL_610758|20 ···················· ···················· ···················· ···················· ···················· ····················

| | | | | | | | | | |
|---|---|---|---|---|---|---|---|---|---|
| O | 3956 | hCoV-19/England/CAMC-A82145/2020\|EPI_ISL_610759\|20 | ···················· | ···················· | ···················· | ···················· | ···················· | ···················· | |
| O | 3957 | hCoV-19/England/CAMC-A81D9D/2020\|EPI_ISL_610760\|20 | ···················· | ···················· | ···················· | ···················· | ···················· | ···················· | |
| O | 3958 | hCoV-19/England/CAMC-A82CEA/2020\|EPI_ISL_610761\|20 | ···················· | ···················· | ···················· | ···················· | ···················· | ···················· | |
| O | 3959 | hCoV-19/England/CAMC-A811FB/2020\|EPI_ISL_610762\|20 | ···················· | ···················· | ···················· | ···················· | ······T············· | ···················· | |
| O | 3960 | hCoV-19/England/CAMC-A82D8D/2020\|EPI_ISL_610763\|20 | ···················· | ···················· | ···················· | ···················· | ···················· | ···················· | |
| O | 3961 | hCoV-19/England/CAMC-A81FB5/2020\|EPI_ISL_610764\|20 | ···················· | ···················· | ···················· | ···················· | ···················· | ···················· | |
| O | 3962 | hCoV-19/England/CAMC-A8242E/2020\|EPI_ISL_610765\|20 | ···················· | ···················· | ···················· | ···················· | ···················· | ···················· | |
| O | 3963 | hCoV-19/England/CAMC-A82233/2020\|EPI_ISL_610767\|20 | ···················· | ···················· | ···················· | ···················· | ···················· | ···················· | |
| O | 3964 | hCoV-19/England/CAMC-A83083/2020\|EPI_ISL_610768\|20 | ···················· | ···················· | ···················· | ···················· | ···················· | ···················· | |
| O | 3965 | hCoV-19/England/CAMC-A8153B/2020\|EPI_ISL_610769\|20 | ···················· | ···················· | ···················· | ···················· | ···················· | ···················· | |
| O | 3966 | hCoV-19/England/CAMC-A81E6D/2020\|EPI_ISL_610770\|20 | ···················· | ···················· | ···················· | ···················· | ···················· | ···················· | |
| O | 3967 | hCoV-19/England/CAMC-A820C0/2020\|EPI_ISL_610772\|20 | ···················· | ···················· | ···················· | ···················· | ···················· | ···················· | |
| O | 3968 | hCoV-19/England/CAMC-A82093/2020\|EPI_ISL_610773\|20 | ···················· | ···················· | ···················· | ···················· | ···················· | ···················· | |
| O | 3969 | hCoV-19/England/CAMC-A8251C/2020\|EPI_ISL_610774\|20 | ···················· | ···················· | ···················· | ···················· | ···················· | ···················· | |
| O | 3970 | hCoV-19/England/CAMC-A80CA0/2020\|EPI_ISL_610775\|20 | ···················· | ···················· | ···················· | ···················· | ···················· | ···················· | |
| O | 3971 | hCoV-19/England/CAMC-A81B75/2020\|EPI_ISL_610776\|20 | ···················· | ···················· | ···················· | ···················· | ···················· | ···················· | |
| O | 3972 | hCoV-19/England/CAMC-A81F1F/2020\|EPI_ISL_610781\|20 | ···················· | ···················· | ···················· | ···················· | ···················· | ···················· | |
| O | 3973 | hCoV-19/England/CAMC-A82989/2020\|EPI_ISL_610782\|20 | ···················· | ···················· | ···················· | ···················· | ···················· | ···················· | |
| O | 3974 | hCoV-19/England/CAMC-A81252/2020\|EPI_ISL_610783\|20 | ···················· | ·A·······A·········· | ···················· | ···················· | ···················· | ···················· | |
| O | 3975 | hCoV-19/England/CAMC-A811DD/2020\|EPI_ISL_610800\|20 | ···················· | ···················· | ···················· | ···················· | ···················· | ···················· | |
| O | 3976 | hCoV-19/England/CAMC-A81E12/2020\|EPI_ISL_610801\|20 | ···················· | ···················· | ···················· | ···················· | ···················· | ···················· | |
| O | 3977 | hCoV-19/England/CAMC-A80D8F/2020\|EPI_ISL_610803\|20 | ···················· | ···················· | ···················· | ···················· | ···················· | ···················· | |
| O | 3978 | hCoV-19/England/CAMC-A82109/2020\|EPI_ISL_610804\|20 | ···················· | ···················· | ···················· | ···················· | ···················· | ···················· | |
| O | 3979 | hCoV-19/England/CAMC-A81F97/2020\|EPI_ISL_610805\|20 | ···················· | ···················· | ···················· | ···················· | ···················· | ···················· | |
| O | 3980 | hCoV-19/England/CAMC-A820EE/2020\|EPI_ISL_610807\|20 | ···················· | ···················· | ···················· | ···················· | ···················· | ···················· | |
| O | 3981 | hCoV-19/England/CAMC-A81DF7/2020\|EPI_ISL_610810\|20 | ···················· | ···················· | ···················· | ···················· | ···················· | ···················· | |
| O | 3982 | hCoV-19/England/CAMC-A829B6/2020\|EPI_ISL_610811\|20 | ···················· | ···················· | ···················· | ···················· | ···················· | ···················· | |
| O | 3983 | hCoV-19/England/CAMC-A810A3/2020\|EPI_ISL_610812\|20 | ···················· | ···················· | ···················· | ···················· | ···················· | ···················· | |
| O | 3984 | hCoV-19/England/CAMC-A81BA2/2020\|EPI_ISL_610813\|20 | ···················· | ···················· | ···················· | ···················· | ···················· | ···················· | |
| O | 3985 | hCoV-19/England/CAMC-A82AB3/2020\|EPI_ISL_610815\|20 | ···················· | ···················· | ···················· | ···················· | ···················· | ···················· | |
| O | 3986 | hCoV-19/England/CAMC-A81F6A/2020\|EPI_ISL_610816\|20 | ···················· | ···················· | ···················· | ···················· | ···················· | ···················· | |
| O | 3987 | hCoV-19/England/CAMC-A82FC3/2020\|EPI_ISL_610817\|20 | ···················· | ···················· | ···················· | ···················· | ···················· | ···················· | |
| O | 3988 | hCoV-19/England/CAMC-A82DF6/2020\|EPI_ISL_610818\|20 | ···················· | ···················· | ···················· | ···················· | ···················· | ···················· | |
| O | 3989 | hCoV-19/England/CAMC-A81F5B/2020\|EPI_ISL_610820\|20 | ···················· | ···················· | ···················· | ···················· | ···················· | ···················· | |
| O | 3990 | hCoV-19/England/CAMC-A82C26/2020\|EPI_ISL_610822\|20 | ···················· | ···················· | ···················· | ···················· | ···················· | ···················· | |
| O | 3991 | hCoV-19/England/CAMC-A81119/2020\|EPI_ISL_610823\|20 | ···················· | ···················· | ···················· | ···················· | ···················· | ···················· | |
| O | 3992 | hCoV-19/England/CAMC-A82DAB/2020\|EPI_ISL_610824\|20 | ···················· | ···················· | ···················· | ···················· | ···················· | ···················· | |
| O | 3993 | hCoV-19/England/CAMC-A80DDA/2020\|EPI_ISL_610825\|20 | ···················· | ·A·················· | ···················· | ···················· | ···················· | ···················· | |
| O | 3994 | hCoV-19/England/CAMC-A829C5/2020\|EPI_ISL_610826\|20 | ···················· | ···················· | ···················· | ···················· | ······T············· | ···················· | |
| O | 3995 | hCoV-19/England/CAMC-A81EB8/2020\|EPI_ISL_610827\|20 | ···················· | ···················· | ···················· | ···················· | ···················· | ···················· | |
| O | 3996 | hCoV-19/England/CAMC-A81E9A/2020\|EPI_ISL_610828\|20 | ···················· | ···················· | ···················· | ···················· | ···················· | ···················· | |
| O | 3997 | hCoV-19/England/CAMC-A8229D/2020\|EPI_ISL_610829\|20 | ···················· | ···················· | ···················· | ···················· | ···················· | ···················· | |
| O | 3998 | hCoV-19/England/CAMC-A82224/2020\|EPI_ISL_610830\|20 | ···················· | ···················· | ···················· | ···················· | ···················· | ···················· | |
| O | 3999 | hCoV-19/England/CAMC-A8102B/2020\|EPI_ISL_610831\|20 | ···················· | ···················· | ···················· | ···················· | ···················· | ···················· | |
| O | 4000 | hCoV-19/Wales/PHWC-480DA3/2020\|EPI_ISL_613000\|2020 | ···················· | ···················· | ···················· | ···················· | ···················· | ···················· | |
| O | 4001 | hCoV-19/Wales/PHWC-480DB2/2020\|EPI_ISL_613001\|2020 | ···················· | ···················· | ···················· | ···················· | ···················· | ···················· | |
| O | 4002 | hCoV-19/Wales/PHWC-480DD0/2020\|EPI_ISL_613003\|2020 | ···················· | ···················· | ···················· | ···················· | ···················· | ···················· | |
| O | 4003 | hCoV-19/Wales/PHWC-480E0A/2020\|EPI_ISL_613006\|2020 | ···················· | ···················· | ···················· | ···················· | ···················· | ···················· | |
| O | 4004 | hCoV-19/Wales/PHWC-480E19/2020\|EPI_ISL_613007\|2020 | ···················· | ···················· | ···················· | ···················· | ···················· | ···················· | |
| O | 4005 | hCoV-19/Wales/PHWC-480E37/2020\|EPI_ISL_613008\|2020 | ···················· | ···················· | ···················· | ···················· | ···················· | ···················· | |
| O | 4006 | hCoV-19/Wales/PHWC-480E64/2020\|EPI_ISL_613009\|2020 | ···················· | ···················· | ···················· | ···················· | ···················· | ···················· | |
| O | 4008 | hCoV-19/Wales/PHWC-480EA0/2020\|EPI_ISL_613011\|2020 | ···················· | ···················· | ···················· | ···················· | ···················· | ···················· | |
| O | 4009 | hCoV-19/Wales/PHWC-480EDD/2020\|EPI_ISL_613012\|2020 | ···················· | ···················· | ···················· | ···················· | ···················· | ···················· | |
| O | 4013 | hCoV-19/Wales/PHWC-480F43/2020\|EPI_ISL_613016\|2020 | ···················· | ···················· | ···················· | ···················· | ···················· | ···················· | |
| O | 4014 | hCoV-19/Wales/PHWC-480F52/2020\|EPI_ISL_613017\|2020 | ···················· | ···················· | ···················· | ···················· | ···················· | ···················· | |
| O | 4015 | hCoV-19/Wales/PHWC-480F61/2020\|EPI_ISL_613018\|2020 | ···················· | ···················· | ···················· | ···················· | ···················· | ···················· | |
| O | 4016 | hCoV-19/Wales/PHWC-480F70/2020\|EPI_ISL_613019\|2020 | ···················· | ···················· | ···················· | ···················· | ···················· | ···················· | |
| O | 4017 | hCoV-19/Wales/PHWC-480F8F/2020\|EPI_ISL_613020\|2020 | ···················· | ···················· | ···················· | ···················· | ···················· | ···················· | |
| O | 4018 | hCoV-19/Wales/PHWC-480F9E/2020\|EPI_ISL_613021\|2020 | ···················· | ···················· | ···················· | ···················· | ···················· | ···················· | |
| O | 4021 | hCoV-19/Wales/PHWC-480FDA/2020\|EPI_ISL_613024\|2020 | ···················· | ···················· | ···················· | ···················· | ···················· | ···················· | |
| O | 4023 | hCoV-19/Wales/PHWC-481003/2020\|EPI_ISL_613026\|2020 | ···················· | ···················· | ···················· | ···················· | ···················· | ···················· | |
| O | 4024 | hCoV-19/Wales/PHWC-481012/2020\|EPI_ISL_613027\|2020 | ···················· | ···················· | ···················· | ···················· | ·NN················· | ··NNN··············· | 1 |
| O | 4025 | hCoV-19/Wales/PHWC-48104F/2020\|EPI_ISL_613028\|2020 | ···················· | ···················· | ···················· | ···················· | ···················· | ···················· | |
| O | 4029 | hCoV-19/Wales/PHWC-4810A9/2020\|EPI_ISL_613032\|2020 | ···················· | ···················· | ···················· | ···················· | ···················· | ···················· | |
| O | 4030 | hCoV-19/Wales/PHWC-4810B8/2020\|EPI_ISL_613033\|2020 | ···················· | ···················· | ···················· | ···················· | ···················· | ···················· | |
| O | 4031 | hCoV-19/Wales/PHWC-4810F4/2020\|EPI_ISL_613034\|2020 | ···················· | ···················· | ···················· | ···················· | ···················· | ···················· | |
| O | 4033 | hCoV-19/Wales/PHWC-48112E/2020\|EPI_ISL_613036\|2020 | ···················· | ···················· | ···················· | ···················· | ···················· | ···················· | |
| O | 4034 | hCoV-19/Wales/PHWC-48113D/2020\|EPI_ISL_613037\|2020 | ···················· | ···················· | ···················· | ···················· | ···················· | ···················· | |
| O | 4035 | hCoV-19/Wales/PHWC-48114C/2020\|EPI_ISL_613038\|2020 | ···················· | ···················· | ···················· | ···················· | ···················· | ···················· | |
| O | 4036 | hCoV-19/Wales/PHWC-481179/2020\|EPI_ISL_613039\|2020 | ···················· | ···················· | ···················· | ···················· | ···················· | ···················· | |
| O | 4037 | hCoV-19/Wales/PHWC-481188/2020\|EPI_ISL_613040\|2020 | ···················· | ···················· | ···················· | ···················· | ···················· | ···················· | |
| O | 4038 | hCoV-19/Wales/PHWC-481197/2020\|EPI_ISL_613041\|2020 | ···················· | ···················· | ···················· | ···················· | ···················· | ···················· | |
| O | 4039 | hCoV-19/Wales/PHWC-4811A6/2020\|EPI_ISL_613042\|2020 | ···················· | ···················· | ···················· | ···················· | ···················· | ···················· | |
| O | 4040 | hCoV-19/Wales/PHWC-4811B5/2020\|EPI_ISL_613043\|2020 | ···················· | ···················· | ···················· | ···················· | ···················· | ···················· | |
| O | 4042 | hCoV-19/Wales/PHWC-48120D/2020\|EPI_ISL_613045\|2020 | ···················· | ···················· | ···················· | ···················· | ···················· | ···················· | |
| O | 4043 | hCoV-19/Wales/PHWC-48123A/2020\|EPI_ISL_613046\|2020 | ···················· | ···················· | ···················· | ···················· | ···················· | ···················· | |
| O | 4044 | hCoV-19/Wales/PHWC-481249/2020\|EPI_ISL_613047\|2020 | ···················· | ···················· | ···················· | ···················· | ···················· | ···················· | |
| O | 4045 | hCoV-19/Wales/PHWC-481267/2020\|EPI_ISL_613048\|2020 | ···················· | ···················· | ···················· | ···················· | ···················· | ···················· | |
| O | 4046 | hCoV-19/Wales/PHWC-481276/2020\|EPI_ISL_613049\|2020 | ···················· | ···················· | ···················· | ···················· | ···················· | ···················· | |
| O | 4048 | hCoV-19/Wales/PHWC-481294/2020\|EPI_ISL_613051\|2020 | ···················· | ···················· | ···················· | ···················· | ···················· | ···················· | |

| | | | | | | | | |
|---|---|---|---|---|---|---|---|---|
| O | 4049 | hCoV-19/Wales/PHWC-4812D0/2020\|EPI_ISL_613052\|2020 | ···················· | ···················· | ····················· | ···················· | ···················· | ···················· |
| O | 4052 | hCoV-19/Wales/PHWC-481319/2020\|EPI_ISL_613055\|2020 | ···················· | ···················· | ····················· | ···················· | ···················· | ···················· |
| O | 4054 | hCoV-19/Wales/PHWC-481346/2020\|EPI_ISL_613057\|2020 | ···················· | ···················· | ····················· | ···················· | ···················· | ···················· |
| O | 4056 | hCoV-19/Wales/PHWC-481364/2020\|EPI_ISL_613059\|2020 | ···················· | ···················· | ····················· | ···················· | ···················· | ···················· |
| O | 4057 | hCoV-19/Wales/PHWC-481373/2020\|EPI_ISL_613060\|2020 | ···················· | ···················· | ····················· | ···················· | ······T············· | ···················· |
| O | 4058 | hCoV-19/Wales/PHWC-481382/2020\|EPI_ISL_613061\|2020 | ···················· | ···················· | ····················· | ···················· | ···················· | ···················· |
| O | 4060 | hCoV-19/Wales/PHWC-4813BF/2020\|EPI_ISL_613063\|2020 | ···················· | ···················· | ····················· | ···················· | ···················· | ···················· |
| O | 4061 | hCoV-19/Wales/PHWC-4813CE/2020\|EPI_ISL_613064\|2020 | ···················· | ···················· | ····················· | ···················· | ···················· | ···················· |
| O | 4062 | hCoV-19/Wales/PHWC-4813DD/2020\|EPI_ISL_613065\|2020 | ···················· | ···················· | ····················· | ···················· | ···················· | ···················· |
| O | 4064 | hCoV-19/Wales/PHWC-481407/2020\|EPI_ISL_613067\|2020 | ···················· | ···················· | ····················· | ···················· | ···················· | ···················· |
| O | 4065 | hCoV-19/Wales/PHWC-481416/2020\|EPI_ISL_613068\|2020 | ···················· | ···················· | ····················· | ···················· | ···················· | ···················· |
| O | 4066 | hCoV-19/Wales/PHWC-481425/2020\|EPI_ISL_613069\|2020 | ···················· | ···················· | ····················· | ···················· | ···················· | ···················· |
| O | 4070 | hCoV-19/Wales/PHWC-48148F/2020\|EPI_ISL_613073\|2020 | ···················· | ···················· | ····················· | ···················· | ···················· | ···················· |
| O | 4072 | hCoV-19/Wales/PHWC-4814AD/2020\|EPI_ISL_613075\|2020 | ···················· | ···················· | ····················· | ···················· | ···················· | ···················· |
| O | 4073 | hCoV-19/Wales/PHWC-4814CB/2020\|EPI_ISL_613076\|2020 | ···················· | ···················· | ····················· | ···················· | ···················· | ···················· |
| O | 4074 | hCoV-19/Wales/PHWC-4814E9/2020\|EPI_ISL_613077\|2020 | ···················· | ···················· | ····················· | ···················· | ···················· | ···················· |
| O | 4076 | hCoV-19/Wales/PHWC-481504/2020\|EPI_ISL_613079\|2020 | ···················· | ···················· | ····················· | ···················· | ···················· | ···················· |
| O | 4077 | hCoV-19/Wales/PHWC-481513/2020\|EPI_ISL_613080\|2020 | ···················· | ···················· | ····················· | ···················· | ···················· | ···················· |
| O | 4078 | hCoV-19/Wales/PHWC-481522/2020\|EPI_ISL_613081\|2020 | ···················· | ···················· | ····················· | ···················· | ···················· | ···················· |
| O | 4079 | hCoV-19/Wales/PHWC-481531/2020\|EPI_ISL_613082\|2020 | ···················· | ···················· | ····················· | ···················· | ···················· | ···················· |
| O | 4080 | hCoV-19/Wales/PHWC-481540/2020\|EPI_ISL_613083\|2020 | ···················· | ···················· | ····················· | ···················· | ···················· | ···················· |
| O | 4081 | hCoV-19/Wales/PHWC-48155F/2020\|EPI_ISL_613084\|2020 | ···················· | ···················· | ····················· | ···················· | ···················· | ···················· |
| O | 4083 | hCoV-19/Wales/PHWC-48157D/2020\|EPI_ISL_613086\|2020 | ···················· | ···················· | ····················· | ···················· | ···················· | ···················· |
| O | 4084 | hCoV-19/Wales/PHWC-48158C/2020\|EPI_ISL_613087\|2020 | ···················· | ···················· | ····················· | ···················· | ···················· | ···················· |
| O | 4085 | hCoV-19/Wales/PHWC-4815AA/2020\|EPI_ISL_613088\|2020 | ···················· | ···················· | ····················· | ···················· | ······T············· | ···················· |
| O | 4086 | hCoV-19/Wales/PHWC-4815B9/2020\|EPI_ISL_613089\|2020 | ···················· | ···················· | ····················· | ···················· | ···················· | ···················· |
| O | 4087 | hCoV-19/Wales/PHWC-4815C8/2020\|EPI_ISL_613090\|2020 | ···················· | ···················· | ····················· | ···················· | ···················· | ···················· |
| O | 4088 | hCoV-19/Wales/PHWC-4815D7/2020\|EPI_ISL_613091\|2020 | ···················· | ···················· | ····················· | ···················· | ···················· | ···················· |
| O | 4089 | hCoV-19/Wales/PHWC-4815E6/2020\|EPI_ISL_613092\|2020 | ···················· | ···················· | ····················· | ···················· | ···················· | ···················· |
| O | 4090 | hCoV-19/Wales/PHWC-4815F5/2020\|EPI_ISL_613093\|2020 | ···················· | ···················· | ····················· | ···················· | ···················· | ···················· |
| O | 4092 | hCoV-19/Wales/PHWC-48163E/2020\|EPI_ISL_613095\|2020 | ···················· | ···················· | ····················· | ···················· | ···················· | ···················· |
| O | 4094 | hCoV-19/Wales/PHWC-48166B/2020\|EPI_ISL_613097\|2020 | ···················· | ···················· | ····················· | ···················· | ···················· | ···················· |
| O | 4096 | hCoV-19/Wales/PHWC-4816A7/2020\|EPI_ISL_613099\|2020 | ···················· | ···················· | ····················· | ···················· | ···················· | ···················· |
| O | 4097 | hCoV-19/England/ALDP-A6FBE6/2020\|EPI_ISL_607100\|20 | ···················· | ···················· | ····················· | ···················· | ···················· | ···················· |
| O | 4098 | hCoV-19/England/ALDP-A6FABC/2020\|EPI_ISL_607101\|20 | ···················· | ···················· | ····················· | ···················· | ···················· | ···················· |
| O | 4099 | hCoV-19/England/ALDP-A6F30C/2020\|EPI_ISL_607102\|20 | ···················· | ···················· | ····················· | ···················· | ···················· | ···················· |
| O | 4100 | hCoV-19/England/ALDP-A6CACE/2020\|EPI_ISL_607103\|20 | ···················· | ···················· | ····················· | ···················· | ···················· | ···················· |
| O | 4101 | hCoV-19/England/ALDP-A6FD1D/2020\|EPI_ISL_607104\|20 | ···················· | ···················· | ····················· | ···················· | ···················· | ···················· |
| O | 4102 | hCoV-19/England/ALDP-A6F7A6/2020\|EPI_ISL_607105\|20 | ···················· | ···················· | ····················· | ···················· | ···················· | ···················· |
| O | 4103 | hCoV-19/England/ALDP-A6EC02/2020\|EPI_ISL_607106\|20 | ···················· | ···················· | ····················· | ···················· | ···················· | ···················· |
| O | 4104 | hCoV-19/England/ALDP-A6E349/2020\|EPI_ISL_607107\|20 | ···················· | ···················· | ····················· | ···················· | ···················· | ···················· |
| O | 4105 | hCoV-19/England/ALDP-A6EE39/2020\|EPI_ISL_607108\|20 | ···················· | ·A·················· | ····················· | ···················· | ···················· | ··A················· |
| O | 4106 | hCoV-19/England/ALDP-A6CC04/2020\|EPI_ISL_607109\|20 | ···················· | ···················· | ····················· | ···················· | ···················· | ···················· |
| O | 4107 | hCoV-19/England/ALDP-A6EE1B/2020\|EPI_ISL_607131\|20 | ···················· | ···················· | ····················· | ···················· | ···················· | ···················· |
| O | 4108 | hCoV-19/England/ALDP-A6D605/2020\|EPI_ISL_607132\|20 | ···················· | ···················· | ····················· | ···················· | ···················· | ···················· |
| O | 4109 | hCoV-19/England/ALDP-A6EB14/2020\|EPI_ISL_607133\|20 | ···················· | ···················· | ····················· | ···················· | ···················· | ···················· |
| O | 4110 | hCoV-19/England/ALDP-A6DE1C/2020\|EPI_ISL_607134\|20 | ···················· | ···················· | ····················· | ···················· | ···················· | ···················· |
| O | 4111 | hCoV-19/England/ALDP-A6D9A2/2020\|EPI_ISL_607135\|20 | ···················· | ···················· | ····················· | ···················· | ······T············· | ···················· |
| O | 4112 | hCoV-19/England/ALDP-A6E288/2020\|EPI_ISL_607136\|20 | ···················· | ···················· | ····················· | ···················· | ···················· | ···················· |
| O | 4113 | hCoV-19/England/ALDP-A6F82B/2020\|EPI_ISL_607137\|20 | ···················· | ···················· | ····················· | ···················· | ···················· | ················A··· |
| O | 4114 | hCoV-19/England/ALDP-A6DD00/2020\|EPI_ISL_607138\|20 | ···················· | ···················· | ····················· | ···················· | ···················· | ···················· |
| O | 4115 | hCoV-19/England/ALDP-A6E4BF/2020\|EPI_ISL_607139\|20 | ···················· | ···················· | ····················· | ···················· | ···················· | ···················· |
| O | 4116 | hCoV-19/England/ALDP-A6E3C1/2020\|EPI_ISL_607140\|20 | ···················· | ·A·······A·········· | ····················· | ···················· | ···················· | ···················· |
| O | 4117 | hCoV-19/England/ALDP-A6F81C/2020\|EPI_ISL_607141\|20 | ···················· | ···················· | ····················· | ···················· | ···················· | ···················· |
| O | 4118 | hCoV-19/England/ALDP-A6E5CB/2020\|EPI_ISL_607142\|20 | ···················· | ···················· | ····················· | ···················· | ···················· | ···················· |
| O | 4119 | hCoV-19/England/ALDP-A6F849/2020\|EPI_ISL_607144\|20 | ···················· | ···················· | ····················· | ···················· | ···················· | ···················· |
| O | 4120 | hCoV-19/England/ALDP-A6EBE7/2020\|EPI_ISL_607145\|20 | ···················· | ···················· | ····················· | ···················· | ···················· | ···················· |
| O | 4121 | hCoV-19/England/ALDP-A6EC11/2020\|EPI_ISL_607146\|20 | ···················· | ···················· | ····················· | ···················· | ···················· | ···················· |
| O | 4122 | hCoV-19/England/ALDP-A6FDB3/2020\|EPI_ISL_607147\|20 | ···················· | ···················· | ····················· | ···················· | ···················· | ···················· |
| O | 4123 | hCoV-19/England/ALDP-A6EA17/2020\|EPI_ISL_607148\|20 | ···················· | ···················· | ····················· | ···················· | ···················· | ···················· |
| O | 4124 | hCoV-19/England/ALDP-A6FD95/2020\|EPI_ISL_607149\|20 | ···················· | ···················· | ····················· | ···················· | ···················· | ···················· |
| O | 4125 | hCoV-19/England/ALDP-A6E6B9/2020\|EPI_ISL_607150\|20 | ···················· | ···················· | ····················· | ···················· | ···················· | ···················· |
| O | 4126 | hCoV-19/England/ALDP-A6F6F4/2020\|EPI_ISL_607151\|20 | ···················· | ···················· | ····················· | ···················· | ···················· | ················A··· |
| O | 4127 | hCoV-19/England/ALDP-A6DD3D/2020\|EPI_ISL_607152\|20 | ···················· | ···················· | ····················· | ···················· | ···················· | ···················· |
| O | 4128 | hCoV-19/England/ALDP-A6E0F7/2020\|EPI_ISL_607153\|20 | ···················· | ···················· | ····················· | ···················· | ···················· | ···················· |
| O | 4129 | hCoV-19/England/ALDP-A6FD3B/2020\|EPI_ISL_607154\|20 | ···················· | ···················· | ····················· | ···················· | ···················· | ···················· |
| O | 4130 | hCoV-19/England/ALDP-A6F2C3/2020\|EPI_ISL_607155\|20 | ···················· | ···················· | ····················· | ···················· | ···················· | ···················· |
| O | 4131 | hCoV-19/England/ALDP-A6E5F8/2020\|EPI_ISL_607156\|20 | ···················· | ···················· | ····················· | ···················· | ···················· | ···················· |
| O | 4132 | hCoV-19/England/ALDP-A6CAEC/2020\|EPI_ISL_607157\|20 | ···················· | ···················· | ····················· | ···················· | ···················· | ···················· |
| O | 4133 | hCoV-19/England/ALDP-A6FCC5/2020\|EPI_ISL_607158\|20 | ···················· | ···················· | ····················· | ···················· | ···················· | ···················· |
| O | 4134 | hCoV-19/England/ALDP-A6D984/2020\|EPI_ISL_607159\|20 | ···················· | ···················· | ····················· | ···················· | ···················· | ···················· |
| O | 4135 | hCoV-19/England/ALDP-A6E2B5/2020\|EPI_ISL_607160\|20 | ···················· | ···················· | ····················· | ···················· | ···················· | ···················· |
| O | 4136 | hCoV-19/England/ALDP-A6FCD4/2020\|EPI_ISL_607161\|20 | ···················· | ···················· | ····················· | ···················· | ···················· | ···················· |
| O | 4137 | hCoV-19/England/ALDP-A6F20F/2020\|EPI_ISL_607162\|20 | ···················· | ···················· | ····················· | ···················· | ···················· | ···················· |
| O | 4138 | hCoV-19/England/ALDP-A6E66E/2020\|EPI_ISL_607163\|20 | ···················· | ···················· | ····················· | ···················· | ···················· | ···················· |
| O | 4139 | hCoV-19/England/ALDP-A6E2D3/2020\|EPI_ISL_607164\|20 | ···················· | ···················· | ····················· | ···················· | ···················· | ···················· |
| O | 4140 | hCoV-19/England/ALDP-A6E15E/2020\|EPI_ISL_607165\|20 | ···················· | ···················· | ····················· | ···················· | ···················· | ···················· |
| O | 4141 | hCoV-19/England/ALDP-A6C74F/2020\|EPI_ISL_607166\|20 | ···················· | ···················· | ····················· | ···················· | ···················· | ···················· |
| O | 4142 | hCoV-19/England/ALDP-A6E2A6/2020\|EPI_ISL_607167\|20 | ···················· | ···················· | ····················· | ···················· | ···················· | ···················· |
| O | 4143 | hCoV-19/England/ALDP-A6F1E4/2020\|EPI_ISL_607168\|20 | ···················· | ···················· | ····················· | ···················· | ···················· | ···················· |

| | | | | | | | | |
|---|---|---|---|---|---|---|---|---|
| O | 4144 | hCoV-19/England/ALDP-A6F928/2020\|EPI_ISL_607170\|20 | ···················· | ···················· | ····················· | ···················· | ···················· | ···················· |
| O | 4145 | hCoV-19/England/ALDP-A6D9B1/2020\|EPI_ISL_607171\|20 | ···················· | ···················· | ····················· | ···················· | ···················· | ···················· |
| O | 4146 | hCoV-19/England/ALDP-A6EEEE/2020\|EPI_ISL_607172\|20 | ···················· | ···················· | ····················· | ···················· | ···················· | ···················· |
| O | 4147 | hCoV-19/England/ALDP-A6E33A/2020\|EPI_ISL_607173\|20 | ···················· | ···················· | ····················· | ···················· | ···················· | ···················· |
| O | 4148 | hCoV-19/England/ALDP-A6E367/2020\|EPI_ISL_607174\|20 | ···················· | ···················· | ····················· | ···················· | ···················· | ···················· |
| O | 4149 | hCoV-19/England/ALDP-A6EB9C/2020\|EPI_ISL_607175\|20 | ···················· | ···················· | ····················· | ···················· | ···················· | ···················· |
| O | 4150 | hCoV-19/England/ALDP-A6E121/2020\|EPI_ISL_607176\|20 | ···················· | ·A·················· | ····················· | ···················· | ···················· | ··A················· |
| O | 4151 | hCoV-19/England/ALDP-A6F72E/2020\|EPI_ISL_607177\|20 | ···················· | ···················· | ····················· | ···················· | ···················· | ···················· |
| O | 4152 | hCoV-19/England/ALDP-A6E385/2020\|EPI_ISL_607178\|20 | ···················· | ···················· | ····················· | ···················· | ···················· | ···················· |
| O | 4153 | hCoV-19/England/ALDP-A6C712/2020\|EPI_ISL_607179\|20 | ···················· | ···················· | ····················· | ···················· | ···················· | ···················· |
| O | 4154 | hCoV-19/England/ALDP-A6EE66/2020\|EPI_ISL_607180\|20 | ···················· | ···················· | ····················· | ···················· | ···················· | ···················· |
| O | 4155 | hCoV-19/England/ALDP-A6F937/2020\|EPI_ISL_607181\|20 | ···················· | ···················· | ····················· | ···················· | ···················· | ···················· |
| O | 4156 | hCoV-19/England/ALDP-A6FC3E/2020\|EPI_ISL_607182\|20 | ···················· | ···················· | ····················· | ···················· | ···················· | ···················· |
| O | 4157 | hCoV-19/England/ALDP-A6ECB7/2020\|EPI_ISL_607183\|20 | ···················· | ···················· | ····················· | ···················· | ···················· | ···················· |
| O | 4158 | hCoV-19/England/ALDP-A6FD2C/2020\|EPI_ISL_607184\|20 | ···················· | ···················· | ····················· | ···················· | ···················· | ···················· |
| O | 4159 | hCoV-19/England/ALDP-A6E604/2020\|EPI_ISL_607185\|20 | ···················· | ···················· | ····················· | ···················· | ···················· | ···················· |
| O | 4160 | hCoV-19/England/ALDP-A6FDE0/2020\|EPI_ISL_607186\|20 | ···················· | ···················· | ····················· | ···················· | ···················· | ···················· |
| O | 4161 | hCoV-19/England/ALDP-A6C994/2020\|EPI_ISL_607187\|20 | ···················· | ···················· | ····················· | ···················· | ···················· | ···················· |
| O | 4162 | hCoV-19/England/ALDP-A6FB8C/2020\|EPI_ISL_607188\|20 | ···················· | ···················· | ····················· | ···················· | ···················· | ···················· |
| O | 4163 | hCoV-19/England/ALDP-A6E3B2/2020\|EPI_ISL_607191\|20 | ···················· | ···················· | ····················· | ···················· | ···················· | ···················· |
| O | 4164 | hCoV-19/England/ALDP-A6E543/2020\|EPI_ISL_607193\|20 | ···················· | ···················· | ···T················· | ···················· | ···················· | ···················· |
| O | 4165 | hCoV-19/England/ALDP-A6ECA8/2020\|EPI_ISL_607194\|20 | ···················· | ···················· | ····················· | ···················· | ···················· | ···················· |
| O | 4166 | hCoV-19/England/ALDP-A6C985/2020\|EPI_ISL_607195\|20 | ···················· | ···················· | ····················· | ···················· | ···················· | ···················· |
| O | 4167 | hCoV-19/England/ALDP-A6CA73/2020\|EPI_ISL_607196\|20 | ···················· | ···················· | ····················· | ···················· | ···················· | ···················· |
| O | 4168 | hCoV-19/England/ALDP-A6CADD/2020\|EPI_ISL_607197\|20 | ···················· | ···················· | ····················· | ···················· | ···················· | ···················· |
| O | 4169 | hCoV-19/England/ALDP-A6EA44/2020\|EPI_ISL_607198\|20 | ···················· | ···················· | ····················· | ···················· | ···················· | ···················· |
| O | 4170 | hCoV-19/England/ALDP-A6F7E2/2020\|EPI_ISL_607199\|20 | ···················· | ···················· | ····················· | ···················· | ···················· | ···················· |
| O | 4171 | hCoV-19/England/CAMC-A65175/2020\|EPI_ISL_608400\|20 | ···················· | ···················· | ····················· | ···················· | ···················· | ···················· |
| O | 4172 | hCoV-19/England/ALDP-A68E4E/2020\|EPI_ISL_608401\|20 | ···················· | ···················· | ····················· | ···················· | ···················· | ···················· |
| O | 4173 | hCoV-19/England/CAMC-A65500/2020\|EPI_ISL_608402\|20 | ···················· | ·A·················· | ····················· | ···················· | ···················· | ···················· |
| O | 4174 | hCoV-19/England/ALDP-A683F4/2020\|EPI_ISL_608403\|20 | ···················· | ·A·················· | ····················· | ···················· | ···················· | ··A················· |
| O | 4175 | hCoV-19/England/ALDP-A6887D/2020\|EPI_ISL_608405\|20 | ···················· | ···················· | ····················· | ···················· | ···················· | ···················· |
| O | 4176 | hCoV-19/England/ALDP-A68C26/2020\|EPI_ISL_608406\|20 | ···················· | ···················· | ····················· | ···················· | ···················· | ···················· |
| O | 4177 | hCoV-19/England/CAMC-A64E06/2020\|EPI_ISL_608407\|20 | ···················· | ···················· | ····················· | ···················· | ···················· | ···················· |
| O | 4178 | hCoV-19/England/CAMC-A6511B/2020\|EPI_ISL_608408\|20 | ···················· | ···················· | ····················· | ···················· | ···················· | ···················· |
| O | 4179 | hCoV-19/England/ALDP-A684B5/2020\|EPI_ISL_608409\|20 | ···················· | ···················· | ····················· | ···················· | ···················· | ···················· |
| O | 4180 | hCoV-19/England/CAMC-A65AB6/2020\|EPI_ISL_608410\|20 | ···················· | ···················· | ····················· | ···················· | ···················· | ···················· |
| O | 4181 | hCoV-19/England/CAMC-A654C7/2020\|EPI_ISL_608411\|20 | ···················· | ···················· | ····················· | ···················· | ···················· | ···················· |
| O | 4182 | hCoV-19/England/CAMC-A65315/2020\|EPI_ISL_608412\|20 | ···················· | ···················· | ····················· | ···················· | ···················· | ···················· |
| O | 4183 | hCoV-19/England/ALDP-A68E99/2020\|EPI_ISL_608413\|20 | ···················· | ···················· | ····················· | ···················· | ···················· | ···················· |
| O | 4184 | hCoV-19/England/ALDP-A67EC7/2020\|EPI_ISL_608414\|20 | ···················· | ···················· | ····················· | ···················· | ···················· | ···················· |
| O | 4185 | hCoV-19/England/ALDP-A685EF/2020\|EPI_ISL_608415\|20 | ···················· | ···················· | ····················· | ···················· | ···················· | ···················· |
| O | 4186 | hCoV-19/England/CAMC-A64B5A/2020\|EPI_ISL_608416\|20 | ···················· | ···················· | ····················· | ···················· | ···················· | ···················· |
| O | 4187 | hCoV-19/England/CAMC-A655B5/2020\|EPI_ISL_608417\|20 | ···················· | ···················· | ····················· | ···················· | ···················· | ···················· |
| O | 4188 | hCoV-19/England/ALDP-A68BA1/2020\|EPI_ISL_608418\|20 | ···················· | ···················· | ····················· | ···················· | ···················· | ···················· |
| O | 4189 | hCoV-19/England/ALDP-A68172/2020\|EPI_ISL_608419\|20 | ···················· | ···················· | ····················· | ···················· | ···················· | ···················· |
| O | 4190 | hCoV-19/England/ALDP-A68BB0/2020\|EPI_ISL_608420\|20 | ···················· | ···················· | ····················· | ···················· | ···················· | ···················· |
| O | 4191 | hCoV-19/England/ALDP-A67DE8/2020\|EPI_ISL_608421\|20 | ···················· | ···················· | ····················· | ···················· | ···················· | ···················· |
| O | 4192 | hCoV-19/England/ALDP-A684F1/2020\|EPI_ISL_608422\|20 | ···················· | ···················· | ····················· | ···················· | ···················· | ···················· |
| O | 4193 | hCoV-19/England/CAMC-A65588/2020\|EPI_ISL_608423\|20 | ···················· | ···················· | ····················· | ···················· | ···················· | ···················· |
| O | 4194 | hCoV-19/England/ALDP-A67DD9/2020\|EPI_ISL_608425\|20 | ···················· | ···················· | ····················· | ···················· | ···················· | ···················· |
| O | 4195 | hCoV-19/England/CAMC-A64D27/2020\|EPI_ISL_608426\|20 | ···················· | ···················· | ····················· | ···················· | ······T············· | ···················· |
| O | 4196 | hCoV-19/England/CAMC-A64C48/2020\|EPI_ISL_608427\|20 | ···················· | ···················· | ····················· | ···················· | ···················· | ···················· |
| O | 4197 | hCoV-19/England/CAMC-A64DAF/2020\|EPI_ISL_608428\|20 | ···················· | ···················· | ····················· | ···················· | ···················· | ···················· |
| O | 4198 | hCoV-19/England/CAMC-A64CDF/2020\|EPI_ISL_608429\|20 | ···················· | ···················· | ····················· | ···················· | ···················· | ···················· |
| O | 4199 | hCoV-19/England/CAMC-A6545E/2020\|EPI_ISL_608432\|20 | ···················· | ···················· | ····················· | ···················· | ···················· | ···················· |
| O | 4200 | hCoV-19/England/CAMC-A65667/2020\|EPI_ISL_608433\|20 | ···················· | ···················· | ····················· | ···················· | ···················· | ···················· |
| O | 4201 | hCoV-19/England/ALDP-A6885F/2020\|EPI_ISL_608434\|20 | ···················· | ···················· | ····················· | ···················· | ···················· | ···················· |
| O | 4202 | hCoV-19/England/ALDP-A68E8A/2020\|EPI_ISL_608435\|20 | ···················· | ···················· | ····················· | ···················· | ···················· | ···················· |
| O | 4203 | hCoV-19/England/CAMC-A655D3/2020\|EPI_ISL_608436\|20 | ···················· | ···················· | ····················· | ···················· | ···················· | ···················· |
| O | 4204 | hCoV-19/England/CAMC-A651B1/2020\|EPI_ISL_608437\|20 | ···················· | ···················· | ····················· | ···················· | ···················· | ···················· |
| O | 4205 | hCoV-19/England/ALDP-A687CB/2020\|EPI_ISL_608438\|20 | ···················· | ···················· | ····················· | ···················· | ···················· | ···················· |
| O | 4206 | hCoV-19/England/ALDP-A6851C/2020\|EPI_ISL_608462\|20 | ···················· | ···················· | ····················· | ···················· | ···················· | ···················· |
| O | 4207 | hCoV-19/England/CAMC-A654B8/2020\|EPI_ISL_608463\|20 | ···················· | ···················· | ····················· | ···················· | ···················· | ···················· |
| O | 4208 | hCoV-19/England/CAMC-A65069/2020\|EPI_ISL_608464\|20 | ···················· | ···················· | ····················· | ···················· | ···················· | ···················· |
| O | 4209 | hCoV-19/England/ALDP-A687DA/2020\|EPI_ISL_608465\|20 | ···················· | ···················· | ····················· | ···················· | ···················· | ···················· |
| O | 4210 | hCoV-19/England/CAMC-A65421/2020\|EPI_ISL_608466\|20 | ···················· | ···················· | ····················· | ···················· | ···················· | ···················· |
| O | 4211 | hCoV-19/England/CAMC-A64F9A/2020\|EPI_ISL_608467\|20 | ···················· | ···················· | ····················· | ···················· | ···················· | ···················· |
| O | 4212 | hCoV-19/England/ALDP-A6845B/2020\|EPI_ISL_608468\|20 | ···················· | ···················· | ····················· | ···················· | ···················· | ···················· |
| O | 4213 | hCoV-19/England/ALDP-A67B57/2020\|EPI_ISL_608469\|20 | ···················· | ···················· | ····················· | ···················· | ···················· | ···················· |
| O | 4214 | hCoV-19/England/ALDP-A683B8/2020\|EPI_ISL_608470\|20 | ···················· | ···················· | ····················· | ···················· | ···················· | ···················· |
| O | 4215 | hCoV-19/England/CAMC-A64D18/2020\|EPI_ISL_608472\|20 | ···················· | ···················· | ····················· | ···················· | ······T············· | ···················· |
| O | 4216 | hCoV-19/England/ALDP-A67E30/2020\|EPI_ISL_608474\|20 | ···················· | ···················· | ····················· | ···················· | ···················· | ···················· |
| O | 4217 | hCoV-19/England/CAMC-A65087/2020\|EPI_ISL_608475\|20 | ···················· | ···················· | ····················· | ···················· | ···················· | ···················· |
| O | 4218 | hCoV-19/England/CAMC-A6561C/2020\|EPI_ISL_608477\|20 | ···················· | ···················· | ····················· | ···················· | ···················· | ···················· |
| O | 4219 | hCoV-19/England/CAMC-A64FA9/2020\|EPI_ISL_608478\|20 | ···················· | ···················· | ····················· | ···················· | ···················· | ···················· |
| O | 4220 | hCoV-19/England/CAMC-A653D9/2020\|EPI_ISL_608480\|20 | ···················· | ···················· | ····················· | ···················· | ···················· | ···················· |
| O | 4221 | hCoV-19/England/ALDP-A68F96/2020\|EPI_ISL_608481\|20 | ···················· | ···················· | ····················· | ···················· | ···················· | ···················· |
| O | 4222 | hCoV-19/England/ALDP-A68E6C/2020\|EPI_ISL_608483\|20 | ···················· | ···················· | ····················· | ···················· | ···················· | ···················· |
| O | 4223 | hCoV-19/England/CAMC-A64D45/2020\|EPI_ISL_608484\|20 | ···················· | ···················· | ····················· | ···················· | ······T············· | ···················· |

| | | | | | | | | |
|---|---|---|---|---|---|---|---|---|
| O | 4224 | hCoV-19/England/CAMC-A64E60/2020\|EPI_ISL_608485\|20 | ···················· | ···················· | ····················· | ···················· | ···················· | ···················· |
| O | 4225 | hCoV-19/England/CAMC-A6595F/2020\|EPI_ISL_608486\|20 | ···················· | ···················· | ····················· | ···················· | ···················· | ···················· |
| O | 4226 | hCoV-19/England/ALDP-A68F78/2020\|EPI_ISL_608487\|20 | ···················· | ···················· | ····················· | ···················· | ···················· | ···················· |
| O | 4227 | hCoV-19/England/ALDP-A68ED5/2020\|EPI_ISL_608488\|20 | ···················· | ···················· | ····················· | ···················· | ···················· | ···················· |
| O | 4228 | hCoV-19/England/ALDP-A68154/2020\|EPI_ISL_608489\|20 | ···················· | ···················· | ····················· | ···················· | ···················· | ···················· |
| O | 4229 | hCoV-19/England/CAMC-A64D36/2020\|EPI_ISL_608490\|20 | ···················· | ···················· | ····················· | ···················· | ···················· | ···················· |
| O | 4230 | hCoV-19/England/ALDP-A68CF9/2020\|EPI_ISL_608491\|20 | ···················· | ···················· | ····················· | ···················· | ···················· | ···················· |
| O | 4231 | hCoV-19/England/CAMC-A64D81/2020\|EPI_ISL_608493\|20 | ···················· | ···················· | ····················· | ···················· | ···················· | ···················· |
| O | 4232 | hCoV-19/England/CAMC-A64DCD/2020\|EPI_ISL_608494\|20 | ···················· | ·A·················· | ····················· | ···················· | ···················· | ··A················· |
| O | 4233 | hCoV-19/England/CAMC-A6547C/2020\|EPI_ISL_608495\|20 | ···················· | ·A·················· | ····················· | ···················· | ···················· | ···················· |
| O | 4234 | hCoV-19/England/ALDP-A688AA/2020\|EPI_ISL_608496\|20 | ···················· | ···················· | ····················· | ···················· | ···················· | ···················· |
| O | 4235 | hCoV-19/England/CAMC-A653AC/2020\|EPI_ISL_608497\|20 | ···················· | ···················· | ····················· | ···················· | ···················· | ···················· |
| O | 4236 | hCoV-19/England/CAMC-A64E42/2020\|EPI_ISL_608498\|20 | ···················· | ···················· | ····················· | ···················· | ···················· | ···················· |
| O | 4237 | hCoV-19/England/ALDP-A685B2/2020\|EPI_ISL_608499\|20 | ···················· | ···················· | ····················· | ···················· | ···················· | ···················· |
| O | 4238 | hCoV-19/England/CAMC-A3E61C/2020\|EPI_ISL_609730\|20 | ···················· | ···················· | ····················· | ···················· | ···················· | ···················· |
| O | 4239 | hCoV-19/England/CAMC-A3F60C/2020\|EPI_ISL_609731\|20 | ···················· | ···················· | ····················· | ···················· | ···················· | ···················· |
| O | 4240 | hCoV-19/England/CAMC-A3F305/2020\|EPI_ISL_609732\|20 | ···················· | ···················· | ····················· | ···················· | ···················· | ···················· |
| O | 4241 | hCoV-19/England/CAMC-A3F851/2020\|EPI_ISL_609734\|20 | ···················· | ···················· | ····················· | ···················· | ···················· | ···················· |
| O | 4242 | hCoV-19/England/CAMC-A3F9E5/2020\|EPI_ISL_609735\|20 | ···················· | ···················· | ····················· | ···················· | ···················· | ···················· |
| O | 4243 | hCoV-19/England/CAMC-A3F6A2/2020\|EPI_ISL_609736\|20 | ···················· | ·A·················· | ····················· | ···················· | ···················· | ···················· |
| O | 4244 | hCoV-19/England/CAMC-A3F912/2020\|EPI_ISL_609737\|20 | ···················· | ···················· | ····················· | ···················· | ···················· | ···················· |
| O | 4245 | hCoV-19/England/CAMC-A3E4C7/2020\|EPI_ISL_609738\|20 | ···················· | ···················· | ····················· | ···················· | ···················· | ···················· |
| O | 4246 | hCoV-19/England/CAMC-A3F727/2020\|EPI_ISL_609740\|20 | ···················· | ···················· | ····················· | ···················· | ···················· | ···················· |
| O | 4247 | hCoV-19/England/CAMC-A3EB4A/2020\|EPI_ISL_609741\|20 | ···················· | ···················· | ····················· | ···················· | ···················· | ···················· |
| O | 4248 | hCoV-19/England/CAMC-A3F47B/2020\|EPI_ISL_609742\|20 | ···················· | ···················· | ····················· | ···················· | ···················· | ···················· |
| O | 4249 | hCoV-19/England/CAMC-A3F675/2020\|EPI_ISL_609743\|20 | ···················· | ···················· | ····················· | ···················· | ···················· | ···················· |
| O | 4250 | hCoV-19/England/CAMC-A3F6DF/2020\|EPI_ISL_609745\|20 | ···················· | ···················· | ····················· | ···················· | ···················· | ···················· |
| O | 4251 | hCoV-19/England/CAMC-A3F806/2020\|EPI_ISL_609747\|20 | ···················· | ···················· | ····················· | ···················· | ···················· | ···················· |
| O | 4252 | hCoV-19/England/CAMC-A3F9C7/2020\|EPI_ISL_609748\|20 | ···················· | ···················· | ····················· | ···················· | ···················· | ···················· |
| O | 4253 | hCoV-19/England/CAMC-A3FA1F/2020\|EPI_ISL_609749\|20 | ···················· | ···················· | ····················· | ···················· | ···················· | ···················· |
| O | 4254 | hCoV-19/England/CAMC-A3EC1A/2020\|EPI_ISL_609750\|20 | ···················· | ···················· | ····················· | ···················· | ···················· | ···················· |
| O | 4255 | hCoV-19/England/CAMC-A3E430/2020\|EPI_ISL_609752\|20 | ···················· | ···················· | ····················· | ···················· | ···················· | ···················· |
| O | 4256 | hCoV-19/England/CAMC-A3EA98/2020\|EPI_ISL_609753\|20 | ···················· | ···················· | ····················· | ···················· | ···················· | ···················· |
| O | 4257 | hCoV-19/England/CAMC-A3EC74/2020\|EPI_ISL_609754\|20 | ···················· | ···················· | ····················· | ···················· | ···················· | ···················· |
| O | 4258 | hCoV-19/England/CAMC-A3EC92/2020\|EPI_ISL_609755\|20 | ···················· | ···················· | ····················· | ···················· | ···················· | ···················· |
| O | 4259 | hCoV-19/England/CAMC-A3F2CC/2020\|EPI_ISL_609756\|20 | ···················· | ···················· | ····················· | ···················· | ···················· | ···················· |
| O | 4260 | hCoV-19/England/CAMC-A3E51F/2020\|EPI_ISL_609757\|20 | ···················· | ·A·················· | ····················· | ···················· | ···················· | ···················· |
| O | 4261 | hCoV-19/England/CAMC-A3F147/2020\|EPI_ISL_609758\|20 | ···················· | ···················· | ····················· | ···················· | ···················· | ···················· |
| O | 4262 | hCoV-19/England/CAMC-A3E55B/2020\|EPI_ISL_609759\|20 | ···················· | ···················· | ····················· | ···················· | ···················· | ···················· |
| O | 4263 | hCoV-19/England/CAMC-A3FB1C/2020\|EPI_ISL_609760\|20 | ···················· | ···················· | ····················· | ···················· | ···················· | ···················· |
| O | 4264 | hCoV-19/England/CAMC-A3F3E7/2020\|EPI_ISL_609761\|20 | ···················· | ···················· | ····················· | ···················· | ···················· | ···················· |
| O | 4265 | hCoV-19/England/CAMC-A3F61B/2020\|EPI_ISL_609762\|20 | ···················· | ···················· | ····················· | ···················· | ···················· | ···················· |
| O | 4266 | hCoV-19/England/CAMC-A3F860/2020\|EPI_ISL_609763\|20 | ···················· | ···················· | ····················· | ···················· | ···················· | ···················· |
| O | 4267 | hCoV-19/England/CAMC-A3F54B/2020\|EPI_ISL_609764\|20 | ···················· | ···················· | ····················· | ···················· | ···················· | ···················· |
| O | 4268 | hCoV-19/England/CAMC-A3E7A0/2020\|EPI_ISL_609765\|20 | ···················· | ···················· | ····················· | ···················· | ···················· | ···················· |
| O | 4269 | hCoV-19/England/CAMC-A3E9B9/2020\|EPI_ISL_609766\|20 | ···················· | ···················· | ····················· | ···················· | ···················· | ···················· |
| O | 4270 | hCoV-19/England/CAMC-A3F50F/2020\|EPI_ISL_609767\|20 | ···················· | ···················· | ····················· | ···················· | ···················· | ···················· |
| O | 4271 | hCoV-19/England/CAMC-A3F648/2020\|EPI_ISL_609769\|20 | ···················· | ···················· | ····················· | ···················· | ···················· | ···················· |
| O | 4272 | hCoV-19/England/CAMC-A3F341/2020\|EPI_ISL_609794\|20 | ···················· | ···················· | ····················· | ···················· | ···················· | ···················· |
| O | 4273 | hCoV-19/England/CAMC-A3E5B5/2020\|EPI_ISL_609795\|20 | ···················· | ···················· | ····················· | ···················· | ···················· | ···················· |
| O | 4274 | hCoV-19/England/CAMC-A3F3D8/2020\|EPI_ISL_609796\|20 | ···················· | ···················· | ····················· | ···················· | ···················· | ···················· |
| O | 4275 | hCoV-19/England/CAMC-A3EC83/2020\|EPI_ISL_609797\|20 | ···················· | ···················· | ····················· | ···················· | ···················· | ···················· |
| O | 4276 | hCoV-19/England/CAMC-A3F0FF/2020\|EPI_ISL_609798\|20 | ···················· | ···················· | ····················· | ···················· | ···················· | ···················· |
| O | 4277 | hCoV-19/England/CAMC-A3E773/2020\|EPI_ISL_609799\|20 | ···················· | ···················· | ····················· | ···················· | ···················· | ···················· |
| O | 4278 | hCoV-19/England/CAMC-A82AE0/2020\|EPI_ISL_610711\|20 | ···················· | ···················· | ····················· | ···················· | ···················· | ···················· |
| O | 4279 | hCoV-19/England/CAMC-A80D52/2020\|EPI_ISL_610712\|20 | ···················· | ···················· | ····················· | ···················· | ···················· | ···················· |
| O | 4280 | hCoV-19/England/CAMC-A80DE9/2020\|EPI_ISL_610714\|20 | ···················· | ···················· | ····················· | ···················· | ······T············· | ···················· |
| O | 4281 | hCoV-19/England/CAMC-A821CD/2020\|EPI_ISL_610715\|20 | ···················· | ···················· | ····················· | ···················· | ···················· | ···················· |
| O | 4282 | hCoV-19/England/CAMC-A83056/2020\|EPI_ISL_610716\|20 | ···················· | ···················· | ····················· | ···················· | ······T············· | ···················· |
| O | 4283 | hCoV-19/England/CAMC-A81A2D/2020\|EPI_ISL_610717\|20 | ···················· | ···················· | ····················· | ···················· | ···················· | ···················· |
| O | 4284 | hCoV-19/England/CAMC-A81A4B/2020\|EPI_ISL_610718\|20 | ···················· | ···················· | ····················· | ···················· | ···················· | ···················· |
| O | 4285 | hCoV-19/England/CAMC-A82F96/2020\|EPI_ISL_610719\|20 | ···················· | ···················· | ····················· | ···················· | ···················· | ···················· |
| O | 4286 | hCoV-19/England/CAMC-A82DC9/2020\|EPI_ISL_610720\|20 | ···················· | ···················· | ····················· | ···················· | ···················· | ···················· |
| O | 4287 | hCoV-19/England/CAMC-A8301A/2020\|EPI_ISL_610721\|20 | ···················· | ···················· | ····················· | ···················· | ······T············· | ···················· |
| O | 4288 | hCoV-19/England/CAMC-A81C09/2020\|EPI_ISL_610722\|20 | ···················· | ·A·················· | ····················· | ···················· | ···················· | ···················· |
| O | 4289 | hCoV-19/England/CAMC-A81067/2020\|EPI_ISL_610723\|20 | ···················· | ···················· | ····················· | ···················· | ···················· | ···················· |
| O | 4290 | hCoV-19/England/CAMC-A81CFA/2020\|EPI_ISL_610724\|20 | ···················· | ···················· | ····················· | ···················· | ···················· | ···················· |
| O | 4291 | hCoV-19/England/CAMC-A81B0C/2020\|EPI_ISL_610725\|20 | ···················· | ···················· | ····················· | ···················· | ···················· | ···················· |
| O | 4292 | hCoV-19/England/CAMC-A8228E/2020\|EPI_ISL_610726\|20 | ···················· | ···················· | ····················· | ···················· | ···················· | ···················· |
| O | 4293 | hCoV-19/England/CAMC-A82118/2020\|EPI_ISL_610728\|20 | ···················· | ···················· | ····················· | ···················· | ···················· | ···················· |
| O | 4294 | hCoV-19/England/CAMC-A82127/2020\|EPI_ISL_610729\|20 | ···················· | ···················· | ····················· | ···················· | ···················· | ···················· |
| O | 4295 | hCoV-19/England/CAMC-A81207/2020\|EPI_ISL_610730\|20 | ···················· | ···················· | ····················· | ···················· | ······T············· | ···················· |
| O | 4296 | hCoV-19/England/CAMC-A81D06/2020\|EPI_ISL_610731\|20 | ···················· | ···················· | ····················· | ···················· | ···················· | ···················· |
| O | 4297 | hCoV-19/England/CAMC-A81137/2020\|EPI_ISL_610732\|20 | ···················· | ···················· | ····················· | ···················· | ···················· | ···················· |
| O | 4298 | hCoV-19/England/CAMC-A8297A/2020\|EPI_ISL_610733\|20 | ···················· | ···················· | ····················· | ···················· | ···················· | ···················· |
| O | 4299 | hCoV-19/England/CAMC-A81234/2020\|EPI_ISL_610734\|20 | ···················· | ···················· | ····················· | ···················· | ······T············· | ···················· |
| O | 4300 | hCoV-19/England/CAMC-A82585/2020\|EPI_ISL_610735\|20 | ···················· | ···················· | ····················· | ···················· | ···················· | ···················· |
| O | 4301 | hCoV-19/England/CAMC-A821AF/2020\|EPI_ISL_610736\|20 | ···················· | ···················· | ····················· | ···················· | ···················· | ···················· |
| O | 4302 | hCoV-19/England/CAMC-A82B1A/2020\|EPI_ISL_610737\|20 | ···················· | ·A·················· | ····················· | ···················· | ···················· | ···················· |
| O | 4303 | hCoV-19/England/CAMC-A820A2/2020\|EPI_ISL_610738\|20 | ···················· | ···················· | ····················· | ···················· | ···················· | ···················· |

| | | | | | | | | |
|---|---|---|---|---|---|---|---|---|
| O | 4304 | hCoV-19/England/CAMC-A82549/2020\|EPI_ISL_610739\|20 | ···················· | ···················· | ····················· | ···················· | ···················· | ···················· |
| O | 4305 | hCoV-19/England/CAMC-A82FB4/2020\|EPI_ISL_610741\|20 | ···················· | ···················· | ····················· | ···················· | ···················· | ···················· |
| O | 4306 | hCoV-19/England/CAMC-A82242/2020\|EPI_ISL_610742\|20 | ···················· | ···················· | ····················· | ···················· | ···················· | ···················· |
| O | 4307 | hCoV-19/England/CAMC-A81BFD/2020\|EPI_ISL_610744\|20 | ···················· | ·R·················· | ····················· | ···················· | ···················· | ···················· |
| O | 4308 | hCoV-19/England/CAMC-A81243/2020\|EPI_ISL_610745\|20 | ···················· | ·A·················· | ····················· | ···················· | ···················· | ···················· |
| O | 4309 | hCoV-19/England/CAMC-A81708/2020\|EPI_ISL_610747\|20 | ···················· | ···················· | ····················· | ···················· | ···················· | ···················· |
| O | 4310 | hCoV-19/England/CAMC-A819F3/2020\|EPI_ISL_610748\|20 | ···················· | ···················· | ····················· | ···················· | ···················· | ···················· |
| O | 4311 | hCoV-19/England/CAMC-A82172/2020\|EPI_ISL_610749\|20 | ···················· | ·A·················· | ····················· | ···················· | ···················· | ···················· |
| O | 4312 | hCoV-19/England/CAMC-A82C9F/2020\|EPI_ISL_610750\|20 | ···················· | ···················· | ····················· | ···················· | ···················· | ···················· |
| O | 4313 | hCoV-19/England/CAMC-A8292F/2020\|EPI_ISL_610751\|20 | ···················· | ···················· | ····················· | ···················· | ···················· | ···················· |
| O | 4314 | hCoV-19/England/CAMC-A81D8E/2020\|EPI_ISL_610752\|20 | ···················· | ···················· | ····················· | ···················· | ···················· | ···················· |
| O | 4315 | hCoV-19/England/CAMC-A82761/2020\|EPI_ISL_610753\|20 | ···················· | ···················· | ····················· | ···················· | ···················· | ···················· |
| O | 4316 | hCoV-19/England/CAMC-A82B0B/2020\|EPI_ISL_610754\|20 | ···················· | ···················· | ····················· | ···················· | ···················· | ···················· |
| O | 4317 | hCoV-19/England/CAMC-A82A95/2020\|EPI_ISL_610777\|20 | ···················· | ···················· | ····················· | ···················· | ···················· | ···················· |
| O | 4318 | hCoV-19/England/CAMC-A82567/2020\|EPI_ISL_610778\|20 | ···················· | ···················· | ····················· | ···················· | ···················· | ···················· |
| O | 4319 | hCoV-19/England/CAMC-A81999/2020\|EPI_ISL_610784\|20 | ···················· | ···················· | ····················· | ···················· | ···················· | ···················· |
| O | 4320 | hCoV-19/England/CAMC-A823C7/2020\|EPI_ISL_610785\|20 | ···················· | ···················· | ····················· | ···················· | ···················· | ···················· |
| O | 4321 | hCoV-19/England/CAMC-A8253A/2020\|EPI_ISL_610786\|20 | ···················· | ·A·················· | ····················· | ···················· | ···················· | ··A················· |
| O | 4322 | hCoV-19/England/CAMC-A8241F/2020\|EPI_ISL_610788\|20 | ···················· | ···················· | ····················· | ···················· | ···················· | ···················· |
| O | 4323 | hCoV-19/England/CAMC-A81717/2020\|EPI_ISL_610789\|20 | ···················· | ···················· | ····················· | ···················· | ···················· | ···················· |
| O | 4324 | hCoV-19/England/CAMC-A81AB4/2020\|EPI_ISL_610790\|20 | ···················· | ···················· | ····················· | ···················· | ···················· | ···················· |
| O | 4325 | hCoV-19/England/CAMC-A81058/2020\|EPI_ISL_610791\|20 | ···················· | ···················· | ····················· | ···················· | ···················· | ···················· |
| O | 4326 | hCoV-19/England/CAMC-A81DE8/2020\|EPI_ISL_610792\|20 | ···················· | ···················· | ····················· | ···················· | ···················· | ···················· |
| O | 4327 | hCoV-19/England/CAMC-A82FD2/2020\|EPI_ISL_610793\|20 | ···················· | ···················· | ····················· | ···················· | ···················· | ···················· |
| O | 4328 | hCoV-19/England/CAMC-A81AA5/2020\|EPI_ISL_610794\|20 | ···················· | ···················· | ····················· | ···················· | ···················· | ···················· |
| O | 4329 | hCoV-19/England/CAMC-A810D0/2020\|EPI_ISL_610795\|20 | ···················· | ·A·················· | ····················· | ···················· | ···················· | ··A················· |
| O | 4330 | hCoV-19/England/CAMC-A823D6/2020\|EPI_ISL_610796\|20 | ···················· | ···················· | ····················· | ···················· | ···················· | ···················· |
| O | 4331 | hCoV-19/England/CAMC-A83065/2020\|EPI_ISL_610797\|20 | ···················· | ···················· | ····················· | ···················· | ···················· | ···················· |
| O | 4332 | hCoV-19/England/CAMC-A81085/2020\|EPI_ISL_610798\|20 | ···················· | ···················· | ····················· | ···················· | ···················· | ···················· |
| O | 4333 | hCoV-19/England/CAMC-A82BFC/2020\|EPI_ISL_610799\|20 | ···················· | ···················· | ····················· | ···················· | ···················· | ···················· |
| O | 4334 | hCoV-19/USA/MI-MDHHS-SC22150/2020\|EPI_ISL_614201\|2 | ···················· | ···················· | ····················· | ···················· | ···················· | ···················· |
| O | 4335 | hCoV-19/USA/MI-MDHHS-SC22152/2020\|EPI_ISL_614203\|2 | ···················· | ···················· | ····················· | ···················· | ···················· | ···················· |
| O | 4336 | hCoV-19/USA/MI-MDHHS-SC22153/2020\|EPI_ISL_614204\|2 | ···················· | ·············A······ | ····················· | ···················· | ···················· | ···················· |
| O | 4337 | hCoV-19/USA/MI-MDHHS-SC22154/2020\|EPI_ISL_614205\|2 | ···················· | ·············A······ | ····················· | ···················· | ···················· | ···················· |
| O | 4338 | hCoV-19/USA/MI-MDHHS-SC22155/2020\|EPI_ISL_614206\|2 | ···················· | ·············A······ | ····················· | ···················· | ···················· | ···················· |
| O | 4339 | hCoV-19/USA/MI-MDHHS-SC22156/2020\|EPI_ISL_614207\|2 | ···················· | ···················· | ····················· | ···················· | ···················· | ···················· |
| O | 4340 | hCoV-19/USA/MI-MDHHS-SC22157/2020\|EPI_ISL_614208\|2 | ···················· | ···················· | ····················· | ···················· | ···················· | ···················· |
| O | 4341 | hCoV-19/USA/MI-MDHHS-SC22158/2020\|EPI_ISL_614209\|2 | ···················· | ···················· | ····················· | ···················· | ···················· | ···················· |
| O | 4342 | hCoV-19/USA/MI-MDHHS-SC22159/2020\|EPI_ISL_614210\|2 | ···················· | ···················· | ····················· | ···················· | ···················· | ···················· |
| O | 4343 | hCoV-19/USA/MI-MDHHS-SC22160/2020\|EPI_ISL_614211\|2 | ···················· | ···················· | ····················· | ···················· | ···················· | ···················· |
| O | 4344 | hCoV-19/USA/MI-MDHHS-SC22161/2020\|EPI_ISL_614212\|2 | ···················· | ···················· | ····················· | ···················· | ···················· | ···················· |
| O | 4345 | hCoV-19/USA/MI-MDHHS-SC22162/2020\|EPI_ISL_614213\|2 | ···················· | ···················· | ····················· | ···················· | ···················· | ···················· |
| O | 4346 | hCoV-19/USA/MI-MDHHS-SC22163/2020\|EPI_ISL_614214\|2 | ···················· | ···················· | ····················· | ···················· | ···················· | ···················· |
| O | 4347 | hCoV-19/USA/MI-MDHHS-SC22164/2020\|EPI_ISL_614215\|2 | ···················· | ·············A······ | ····················· | ···················· | ···················· | ···················· |
| O | 4348 | hCoV-19/USA/MI-MDHHS-SC22165/2020\|EPI_ISL_614216\|2 | ···················· | ·A···A·············· | ····················· | ···················· | ···················· | ···················· |
| O | 4349 | hCoV-19/USA/MI-MDHHS-SC22166/2020\|EPI_ISL_614217\|2 | ···················· | ···················· | ····················· | ···················· | ···················· | ···················· |
| O | 4350 | hCoV-19/USA/MI-MDHHS-SC22167/2020\|EPI_ISL_614218\|2 | ···················· | ···················· | ····················· | ···················· | ···················· | ···················· |
| O | 4351 | hCoV-19/USA/MI-MDHHS-SC22168/2020\|EPI_ISL_614219\|2 | ···················· | ···················· | ····················· | ···················· | ···················· | ···················· |
| O | 4352 | hCoV-19/USA/MI-MDHHS-SC22175/2020\|EPI_ISL_614226\|2 | ···················· | ···················· | ····················· | ···················· | ···················· | ···················· |
| O | 4353 | hCoV-19/England/ALDP-A6D571/2020\|EPI_ISL_607000\|20 | ···················· | ···················· | ····················· | ···················· | ···················· | ···················· |
| O | 4354 | hCoV-19/England/ALDP-A6DD4C/2020\|EPI_ISL_607001\|20 | ···················· | ···················· | ····················· | ···················· | ···················· | ···················· |
| O | 4355 | hCoV-19/England/ALDP-A6CA19/2020\|EPI_ISL_607002\|20 | ···················· | ···················· | ····················· | ···················· | ···················· | ···················· |
| O | 4356 | hCoV-19/England/ALDP-A6E19A/2020\|EPI_ISL_607003\|20 | ···················· | ···················· | ····················· | ···················· | ···················· | ···················· |
| O | 4357 | hCoV-19/England/ALDP-A6D553/2020\|EPI_ISL_607004\|20 | ···················· | ···················· | ····················· | ···················· | ···················· | ···················· |
| O | 4358 | hCoV-19/England/ALDP-A6DE2B/2020\|EPI_ISL_607005\|20 | ···················· | ···················· | ····················· | ···················· | ···················· | ···················· |
| O | 4359 | hCoV-19/England/ALDP-A6EC7B/2020\|EPI_ISL_607006\|20 | ···················· | ···················· | ····················· | ···················· | ···················· | ···················· |
| O | 4360 | hCoV-19/England/ALDP-A6EBD8/2020\|EPI_ISL_607007\|20 | ···················· | ···················· | ····················· | ···················· | ···················· | ···················· |
| O | 4361 | hCoV-19/England/ALDP-A6F867/2020\|EPI_ISL_607008\|20 | ···················· | ···················· | ····················· | ···················· | ···················· | ···················· |
| O | 4362 | hCoV-19/England/ALDP-A6F2A5/2020\|EPI_ISL_607009\|20 | ···················· | ···················· | ····················· | ···················· | ···················· | ···················· |
| O | 4363 | hCoV-19/England/ALDP-A6EC3F/2020\|EPI_ISL_607010\|20 | ···················· | ···················· | ····················· | ···················· | ···················· | ···················· |
| O | 4364 | hCoV-19/England/ALDP-A6E2E2/2020\|EPI_ISL_607011\|20 | ···················· | ···················· | ····················· | ···················· | ···················· | ···················· |
| O | 4365 | hCoV-19/England/ALDP-A6EE2A/2020\|EPI_ISL_607012\|20 | ···················· | ···················· | ····················· | ···················· | ···················· | ···················· |
| O | 4366 | hCoV-19/England/ALDP-A6E21F/2020\|EPI_ISL_607013\|20 | ···················· | ···················· | ····················· | ···················· | ···················· | ···················· |
| O | 4367 | hCoV-19/England/ALDP-A6E2C4/2020\|EPI_ISL_607014\|20 | ···················· | ···················· | ····················· | ···················· | ···················· | ···················· |
| O | 4368 | hCoV-19/England/ALDP-A6F014/2020\|EPI_ISL_607015\|20 | ···················· | ···················· | ····················· | ···················· | ···················· | ···················· |
| O | 4369 | hCoV-19/England/ALDP-A6C967/2020\|EPI_ISL_607016\|20 | ···················· | ···················· | ····················· | ···················· | ···················· | ···················· |
| O | 4370 | hCoV-19/England/ALDP-A6DE0D/2020\|EPI_ISL_607017\|20 | ···················· | ···················· | ····················· | ···················· | ···················· | ···················· |
| O | 4371 | hCoV-19/England/ALDP-A6EE0C/2020\|EPI_ISL_607018\|20 | ···················· | ···················· | ····················· | ···················· | ···················· | ···················· |
| O | 4372 | hCoV-19/England/ALDP-A6EE57/2020\|EPI_ISL_607019\|20 | ···················· | ···················· | ····················· | ···················· | ···················· | ···················· |
| O | 4373 | hCoV-19/England/ALDP-A6C79A/2020\|EPI_ISL_607020\|20 | ···················· | ···················· | ····················· | ···················· | ···················· | ···················· |
| O | 4374 | hCoV-19/England/ALDP-A6FC98/2020\|EPI_ISL_607021\|20 | ···················· | ···················· | ····················· | ···················· | ···················· | ···················· |
| O | 4375 | hCoV-19/England/ALDP-A6C949/2020\|EPI_ISL_607023\|20 | ···················· | ·A·················· | ····················· | ···················· | ···················· | ··A················· |
| O | 4376 | hCoV-19/England/ALDP-A6FB22/2020\|EPI_ISL_607024\|20 | ···················· | ···················· | ····················· | ···················· | ···················· | ···················· |
| O | 4377 | hCoV-19/England/ALDP-A6E1A9/2020\|EPI_ISL_607025\|20 | ···················· | ·A·················· | ····················· | ···················· | ···················· | ··A················· |
| O | 4378 | hCoV-19/England/ALDP-A6CC31/2020\|EPI_ISL_607026\|20 | ···················· | ···················· | ····················· | ···················· | ···················· | ···················· |
| O | 4379 | hCoV-19/England/ALDP-A6FDC2/2020\|EPI_ISL_607028\|20 | ···················· | ···················· | ····················· | ···················· | ···················· | ···················· |
| O | 4380 | hCoV-19/England/ALDP-A6DDF1/2020\|EPI_ISL_607029\|20 | ···················· | ···················· | ····················· | ···················· | ···················· | ···················· |
| O | 4381 | hCoV-19/England/ALDP-A6FE74/2020\|EPI_ISL_608320\|20 | ···················· | ···················· | ····················· | ···················· | ···················· | ···················· |
| O | 4382 | hCoV-19/England/ALDP-A70881/2020\|EPI_ISL_608321\|20 | ···················· | ···················· | ····················· | ···················· | ···················· | ···················· |
| O | 4383 | hCoV-19/England/ALDP-A6CD3E/2020\|EPI_ISL_608323\|20 | ···················· | ···················· | ····················· | ···················· | ···················· | ···················· |

| | | | | | | | | |
|---|---|---|---|---|---|---|---|---|
| O | 4384 | hCoV-19/England/ALDP-A6D034/2020\|EPI_ISL_608324\|20 | .................... | .................... | .................... | .................... | .................... | .................... |
| O | 4385 | hCoV-19/England/ALDP-A6C6BB/2020\|EPI_ISL_608325\|20 | .................... | .................... | .................... | .................... | .................... | .................... |
| O | 4386 | hCoV-19/England/ALDP-A6C3A5/2020\|EPI_ISL_608326\|20 | .................... | .................... | .................... | .................... | .................... | .................... |
| O | 4387 | hCoV-19/England/ALDP-A70845/2020\|EPI_ISL_608327\|20 | .................... | .................... | .................... | .................... | .................... | .................... |
| O | 4388 | hCoV-19/England/ALDP-A6C4FD/2020\|EPI_ISL_608328\|20 | .................... | .................... | .................... | .................... | .................... | .................... |
| O | 4389 | hCoV-19/England/ALDP-A6C396/2020\|EPI_ISL_608329\|20 | .................... | .................... | .................... | .................... | ......T............. | .................... |
| O | 4390 | hCoV-19/England/ALDP-A7021A/2020\|EPI_ISL_608330\|20 | .................... | .................... | .................... | .................... | .................... | .................... |
| O | 4391 | hCoV-19/England/ALDP-A6C509/2020\|EPI_ISL_608331\|20 | .................... | .................... | .................... | .................... | .................... | .................... |
| O | 4392 | hCoV-19/England/ALDP-A6C41B/2020\|EPI_ISL_608332\|20 | .................... | .A.................. | .................... | .................... | .................... | .................... |
| O | 4393 | hCoV-19/England/ALDP-A6FE65/2020\|EPI_ISL_608333\|20 | .................... | .................... | .................... | .................... | .................... | .................... |
| O | 4394 | hCoV-19/England/ALDP-A7013B/2020\|EPI_ISL_608334\|20 | .................... | .................... | .................... | .................... | .................... | .................... |
| O | 4395 | hCoV-19/England/ALDP-A6D1B9/2020\|EPI_ISL_608335\|20 | .................... | .................... | .................... | .................... | .................... | .................... |
| O | 4396 | hCoV-19/England/ALDP-A701E0/2020\|EPI_ISL_608336\|20 | .................... | .................... | .................... | .................... | .................... | .................... |
| O | 4397 | hCoV-19/England/ALDP-A6C84C/2020\|EPI_ISL_608337\|20 | .................... | .................... | .................... | .................... | .................... | .................... |
| O | 4398 | hCoV-19/England/ALDP-A6FEDE/2020\|EPI_ISL_608338\|20 | .................... | .................... | .................... | .................... | .................... | .................... |
| O | 4399 | hCoV-19/England/ALDP-A6C5EB/2020\|EPI_ISL_608339\|20 | .................... | .................... | .................... | .................... | .................... | .................... |
| O | 4400 | hCoV-19/England/ALDP-A6D007/2020\|EPI_ISL_608340\|20 | .................... | .................... | .................... | .................... | .................... | .................... |
| O | 4401 | hCoV-19/England/ALDP-A6FFEA/2020\|EPI_ISL_608341\|20 | .................... | .................... | .................... | .................... | .................... | .................... |
| O | 4402 | hCoV-19/England/ALDP-A6C35A/2020\|EPI_ISL_608342\|20 | .................... | .................... | .................... | .................... | .................... | .................... |
| O | 4403 | hCoV-19/England/ALDP-A6D68D/2020\|EPI_ISL_608343\|20 | .................... | .................... | .................... | .................... | .................... | .................... |
| O | 4404 | hCoV-19/England/ALDP-A6FFF9/2020\|EPI_ISL_608344\|20 | .................... | .................... | .................... | .................... | .................... | .................... |
| O | 4405 | hCoV-19/England/ALDP-A6C90D/2020\|EPI_ISL_608345\|20 | .................... | .................... | .................... | .................... | .................... | .................... |
| O | 4406 | hCoV-19/England/CAMC-A64F8B/2020\|EPI_ISL_608348\|20 | .................... | .................... | .................... | .................... | .................... | .................... |
| O | 4407 | hCoV-19/England/CAMC-A65148/2020\|EPI_ISL_608349\|20 | .................... | .................... | .................... | .................... | .................... | .................... |
| O | 4408 | hCoV-19/England/ALDP-A688C8/2020\|EPI_ISL_608350\|20 | .................... | .................... | .................... | .................... | .................... | .................... |
| O | 4409 | hCoV-19/England/ALDP-A68215/2020\|EPI_ISL_608351\|20 | .................... | .................... | .................... | .................... | .................... | .................... |
| O | 4410 | hCoV-19/England/ALDP-A68FA5/2020\|EPI_ISL_608352\|20 | .................... | .................... | .................... | .................... | .................... | .................... |
| O | 4411 | hCoV-19/England/CAMC-A658E9/2020\|EPI_ISL_608353\|20 | .................... | .A.................. | .................... | .................... | .................... | .................... |
| O | 4412 | hCoV-19/England/CAMC-A6546D/2020\|EPI_ISL_608354\|20 | .................... | .................... | .................... | .................... | .................... | .................... |
| O | 4413 | hCoV-19/England/ALDP-A6844C/2020\|EPI_ISL_608355\|20 | .................... | .................... | .................... | .................... | .................... | .................... |
| O | 4414 | hCoV-19/England/ALDP-A68CEA/2020\|EPI_ISL_608356\|20 | .................... | .................... | .................... | .................... | .................... | .................... |
| O | 4415 | hCoV-19/England/CAMC-A64F4F/2020\|EPI_ISL_608357\|20 | .................... | .................... | .................... | .................... | .................... | .................... |
| O | 4416 | hCoV-19/England/ALDP-A67F3D/2020\|EPI_ISL_608358\|20 | .................... | .................... | .................... | .................... | .................... | .................... |
| O | 4417 | hCoV-19/England/CAMC-A65403/2020\|EPI_ISL_608359\|20 | .................... | .................... | .................... | .................... | .................... | .................... |
| O | 4418 | hCoV-19/England/CAMC-A3F842/2020\|EPI_ISL_609650\|20 | .................... | .................... | .................... | .................... | .................... | .................... |
| O | 4419 | hCoV-19/England/CAMC-A3E4B8/2020\|EPI_ISL_609651\|20 | .................... | .................... | .................... | .................... | .................... | .................... |
| O | 4420 | hCoV-19/England/CAMC-A3E70A/2020\|EPI_ISL_609652\|20 | .................... | .................... | .................... | .................... | .................... | .................... |
| O | 4421 | hCoV-19/England/CAMC-A3F1FC/2020\|EPI_ISL_609653\|20 | .................... | .................... | .................... | .................... | .................... | .................... |
| O | 4422 | hCoV-19/England/CAMC-A3E5F1/2020\|EPI_ISL_609654\|20 | .................... | .................... | .................... | .................... | .................... | .................... |
| O | 4423 | hCoV-19/England/CAMC-A3ECDE/2020\|EPI_ISL_609655\|20 | .................... | .................... | .................... | .................... | .................... | .................... |
| O | 4424 | hCoV-19/England/CAMC-A3E8CB/2020\|EPI_ISL_609657\|20 | .................... | .................... | .................... | .................... | .................... | .................... |
| O | 4425 | hCoV-19/England/CAMC-A3EB86/2020\|EPI_ISL_609659\|20 | .................... | .................... | .................... | .................... | .................... | .................... |
| O | 4426 | hCoV-19/England/CAMC-A3E500/2020\|EPI_ISL_609660\|20 | .................... | .................... | .................... | .................... | .................... | .................... |
| O | 4427 | hCoV-19/England/CAMC-A3FA5B/2020\|EPI_ISL_609661\|20 | .................... | .................... | .................... | .................... | .................... | .................... |
| O | 4428 | hCoV-19/England/CAMC-A3F781/2020\|EPI_ISL_609662\|20 | .................... | .................... | .................... | .................... | .................... | .................... |
| O | 4429 | hCoV-19/England/CAMC-A3F0B3/2020\|EPI_ISL_609665\|20 | .................... | .................... | .................... | .................... | ......T............. | .................... |
| O | 4430 | hCoV-19/England/CAMC-A3EA5C/2020\|EPI_ISL_609666\|20 | .................... | .................... | .................... | .................... | .................... | .................... |
| O | 4431 | hCoV-19/England/CAMC-A3F97C/2020\|EPI_ISL_609667\|20 | .................... | .................... | .................... | .................... | .................... | .................... |
| O | 4432 | hCoV-19/England/CAMC-A3E4E5/2020\|EPI_ISL_609668\|20 | .................... | .................... | .................... | .................... | .................... | .................... |
| O | 4433 | hCoV-19/England/CAMC-A3EA7A/2020\|EPI_ISL_609669\|20 | .................... | .................... | .................... | .................... | ......T............. | .................... |
| O | 4434 | hCoV-19/England/CAMC-A3F499/2020\|EPI_ISL_609670\|20 | .................... | .................... | .................... | .................... | .................... | .................... |
| O | 4435 | hCoV-19/England/CAMC-A3F11A/2020\|EPI_ISL_609671\|20 | .................... | .A.................. | .................... | .................... | .................... | ...A................ |
| O | 4436 | hCoV-19/England/CAMC-A3EE50/2020\|EPI_ISL_609673\|20 | .................... | .................... | .................... | .................... | .................... | .................... |
| O | 4437 | hCoV-19/England/CAMC-A3F98B/2020\|EPI_ISL_609674\|20 | .................... | .................... | .................... | .................... | .................... | .................... |
| O | 4438 | hCoV-19/England/CAMC-A3EB0E/2020\|EPI_ISL_609675\|20 | .................... | .................... | .................... | .................... | .................... | .................... |
| O | 4439 | hCoV-19/England/CAMC-A3EF8A/2020\|EPI_ISL_609676\|20 | .................... | .................... | .................... | .................... | .................... | .................... |
| O | 4440 | hCoV-19/England/CAMC-A3E98C/2020\|EPI_ISL_609678\|20 | .................... | .................... | .................... | .................... | .................... | .................... |
| O | 4441 | hCoV-19/England/CAMC-A3F903/2020\|EPI_ISL_609679\|20 | .................... | .................... | .................... | .................... | .................... | .................... |
| O | 4442 | hCoV-19/England/CAMC-A3E3D9/2020\|EPI_ISL_609680\|20 | .................... | .................... | .................... | .................... | .................... | .................... |
| O | 4443 | hCoV-19/England/CAMC-A3F38D/2020\|EPI_ISL_609681\|20 | .................... | .................... | .................... | .................... | .................... | .................... |
| O | 4444 | hCoV-19/England/CAMC-A3F96D/2020\|EPI_ISL_609682\|20 | .................... | .................... | .................... | .................... | .................... | .................... |
| O | 4445 | hCoV-19/England/CAMC-A3F5E1/2020\|EPI_ISL_609683\|20 | .................... | .................... | .................... | .................... | .................... | .................... |
| O | 4446 | hCoV-19/England/CAMC-A3FB94/2020\|EPI_ISL_609684\|20 | .................... | .................... | .................... | .................... | .................... | .................... |
| O | 4447 | hCoV-19/England/CAMC-A3E4A9/2020\|EPI_ISL_609685\|20 | .................... | .................... | .................... | .................... | .................... | .................... |
| O | 4448 | hCoV-19/England/CAMC-A3F2F9/2020\|EPI_ISL_609686\|20 | .................... | .................... | .................... | .................... | .................... | .................... |
| O | 4449 | hCoV-19/England/CAMC-A3EDDB/2020\|EPI_ISL_609687\|20 | .................... | .................... | .................... | .................... | .................... | .................... |
| O | 4450 | hCoV-19/England/CAMC-A3F208/2020\|EPI_ISL_609688\|20 | .................... | .................... | .................... | .................... | .................... | .................... |
| O | 4451 | hCoV-19/England/CAMC-A3E52E/2020\|EPI_ISL_609689\|20 | .................... | .................... | .................... | .................... | .................... | .................... |
| O | 4452 | hCoV-19/England/CAMC-A3E46D/2020\|EPI_ISL_609700\|20 | .................... | .................... | .................... | .................... | .................... | .................... |
| O | 4453 | hCoV-19/England/CAMC-A3E9E6/2020\|EPI_ISL_609702\|20 | .................... | .................... | .................... | .................... | .................... | .................... |
| O | 4454 | hCoV-19/England/CAMC-A3F077/2020\|EPI_ISL_609704\|20 | .................... | .................... | .................... | .................... | .................... | .................... |
| O | 4455 | hCoV-19/England/CAMC-A3FB2B/2020\|EPI_ISL_609705\|20 | .................... | .................... | .................... | .................... | .................... | .................... |
| O | 4456 | hCoV-19/England/CAMC-A3F10B/2020\|EPI_ISL_609706\|20 | .................... | .................... | .................... | .................... | .................... | .................... |
| O | 4457 | hCoV-19/England/CAMC-A3EC0B/2020\|EPI_ISL_609707\|20 | .................... | .A.................. | .................... | .................... | .................... | ...A................ |
| O | 4458 | hCoV-19/England/CAMC-A3F37E/2020\|EPI_ISL_609708\|20 | .................... | .................... | .................... | .................... | .................... | .................... |
| O | 4459 | hCoV-19/England/CAMC-A3ECED/2020\|EPI_ISL_609709\|20 | .................... | .................... | .................... | .................... | .................... | .................... |
| O | 4460 | hCoV-19/England/CAMC-A3E8E9/2020\|EPI_ISL_609710\|20 | .................... | .................... | .................... | .................... | .................... | .................... |
| O | 4461 | hCoV-19/England/CAMC-A3F815/2020\|EPI_ISL_609711\|20 | .................... | .................... | .................... | .................... | .................... | .................... |
| O | 4462 | hCoV-19/England/CAMC-A3F2DB/2020\|EPI_ISL_609712\|20 | .................... | .................... | .................... | .................... | .................... | .................... |
| O | 4463 | hCoV-19/England/CAMC-A3E931/2020\|EPI_ISL_609713\|20 | .................... | .................... | .................... | .................... | .................... | .................... |

O 4464 hCoV-19/England/CAMC-A3F7DC/2020|EPI_ISL_609714|20 ···················· ···················· ····················· ···················· ···················· ····················
O 4465 hCoV-19/England/CAMC-A3F04A/2020|EPI_ISL_609715|20 ···················· ···················· ····················· ···················· ···················· ····················
O 4466 hCoV-19/England/CAMC-A3F4F3/2020|EPI_ISL_609716|20 ···················· ···················· ····················· ···················· ···················· ····················
O 4467 hCoV-19/England/CAMC-A3E685/2020|EPI_ISL_609717|20 ···················· ···················· ····················· ···················· ···················· ····················
O 4468 hCoV-19/England/CAMC-A3F068/2020|EPI_ISL_609718|20 ···················· ···················· ····················· ···················· ···················· ····················
O 4469 hCoV-19/England/CAMC-A3E3F7/2020|EPI_ISL_609719|20 ···················· ···················· ····················· ···················· ···················· ····················
O 4470 hCoV-19/England/CAMC-A3FA00/2020|EPI_ISL_609720|20 ···················· ···················· ····················· ···················· ···················· ····················
O 4471 hCoV-19/England/CAMC-A3F8D9/2020|EPI_ISL_609721|20 ···················· ···················· ····················· ···················· ···················· ····················
O 4472 hCoV-19/England/CAMC-A3F51E/2020|EPI_ISL_609722|20 ···················· ···················· ····················· ···················· ···················· ····················
O 4473 hCoV-19/England/CAMC-A3E421/2020|EPI_ISL_609723|20 ···················· ···················· ····················· ···················· ···················· ····················
O 4474 hCoV-19/England/CAMC-A3F235/2020|EPI_ISL_609724|20 ···················· ···················· ····················· ···················· ···················· ····················
O 4475 hCoV-19/England/CAMC-A3F639/2020|EPI_ISL_609725|20 ···················· ···················· ····················· ···················· ······T············· ····················
O 4476 hCoV-19/England/CAMC-A3EAE3/2020|EPI_ISL_609726|20 ···················· ···················· ····················· ···················· ···················· ····················
O 4477 hCoV-19/England/CAMC-A3E47C/2020|EPI_ISL_609727|20 ···················· ···················· ····················· ···················· ···················· ····················
O 4478 hCoV-19/England/CAMC-A3FB85/2020|EPI_ISL_609728|20 ···················· ···················· ····················· ···················· ···················· ····················
O 4479 hCoV-19/England/CAMC-A3F666/2020|EPI_ISL_609729|20 ···················· ···················· ····················· ···················· ···················· ····················
O 4480 hCoV-19/England/CAMC-A80D61/2020|EPI_ISL_610634|20 ···················· ···················· ····················· ···················· ···················· ····················
O 4481 hCoV-19/England/CAMC-A81304/2020|EPI_ISL_610635|20 ···················· ···················· ····················· ···················· ···················· ····················
O 4482 hCoV-19/England/CAMC-A8101C/2020|EPI_ISL_610636|20 ···················· ···················· ····················· ···················· ···················· ····················
O 4483 hCoV-19/England/CAMC-A81155/2020|EPI_ISL_610637|20 ···················· ···················· ····················· ···················· ···················· ····················
O 4484 hCoV-19/England/CAMC-A82576/2020|EPI_ISL_610638|20 ···················· ···················· ····················· ···················· ···················· ····················
O 4485 hCoV-19/England/CAMC-A81225/2020|EPI_ISL_610640|20 ···················· ···················· ····················· ···················· ······T············· ····················
O 4486 hCoV-19/Wales/CAMC-A80D43/2020|EPI_ISL_610641|2020 ···················· ···················· ····················· ···················· ···················· ····················
O 4487 hCoV-19/England/CAMC-A80DCB/2020|EPI_ISL_610642|20 ···················· ···················· ····················· ···················· ···················· ····················
O 4488 hCoV-19/England/CAMC-A82594/2020|EPI_ISL_610643|20 ···················· ···················· ····················· ···················· ···················· ····················
O 4489 hCoV-19/England/CAMC-A8201B/2020|EPI_ISL_610644|20 ···················· ···················· ····················· ···················· ···················· ····················
O 4490 hCoV-19/England/CAMC-A82C35/2020|EPI_ISL_610645|20 ···················· ·A·······A·········· ····················· ···················· ···················· ····················
O 4491 hCoV-19/England/CAMC-A82CCC/2020|EPI_ISL_610646|20 ···················· ···················· ····················· ···················· ···················· ····················
O 4492 hCoV-19/England/CAMC-A8103A/2020|EPI_ISL_610647|20 ···················· ···················· ····················· ···················· ···················· ····················
O 4493 hCoV-19/England/CAMC-A81A3C/2020|EPI_ISL_610649|20 ···················· ···················· ····················· ···················· ···················· ····················
O 4494 hCoV-19/England/CAMC-A824F1/2020|EPI_ISL_610650|20 ···················· ···················· ····················· ···················· ···················· ····················
O 4495 hCoV-19/England/CAMC-A821BE/2020|EPI_ISL_610651|20 ···················· ···················· ····················· ···················· ···················· ····················
O 4496 hCoV-19/England/CAMC-A81B57/2020|EPI_ISL_610652|20 ···················· ···················· ····················· ···················· ···················· ····················
O 4497 hCoV-19/England/CAMC-A81C63/2020|EPI_ISL_610653|20 ···················· ···················· ····················· ···················· ···················· ····················
O 4498 hCoV-19/England/CAMC-A8196C/2020|EPI_ISL_610655|20 ···················· ···················· ····················· ···················· ···················· ····················
O 4499 hCoV-19/England/CAMC-A81A69/2020|EPI_ISL_610656|20 ···················· ···················· ····················· ···················· ···················· ····················
O 4500 hCoV-19/England/CAMC-A829D4/2020|EPI_ISL_610657|20 ···················· ···················· ····················· ···················· ···················· ····················
O 4501 hCoV-19/England/CAMC-A82E5D/2020|EPI_ISL_610658|20 ···················· ···················· ····················· ···················· ···················· ····················
O 4502 hCoV-19/England/CAMC-A82C53/2020|EPI_ISL_610659|20 ···················· ·A·················· ····················· ···················· ···················· ····················
O 4503 hCoV-19/England/CAMC-A81076/2020|EPI_ISL_610660|20 ···················· ···················· ····················· ···················· ···················· ····················
O 4504 hCoV-19/England/CAMC-A82C17/2020|EPI_ISL_610661|20 ···················· ·A·················· ····················· ···················· ···················· ···A················
O 4505 hCoV-19/England/CAMC-A81FF1/2020|EPI_ISL_610662|20 ···················· ···················· ····················· ···················· ···················· ····················
O 4506 hCoV-19/England/CAMC-A82DE7/2020|EPI_ISL_610663|20 ···················· ·A·················· ····················· ···················· ···················· ···A················
O 4507 hCoV-19/England/CAMC-A81A5A/2020|EPI_ISL_610664|20 ···················· ···················· ····················· ···················· ···················· ····················
O 4508 hCoV-19/England/CAMC-A83092/2020|EPI_ISL_610665|20 ···················· ·A·················· ····················· ···················· ···················· ····················
O 4509 hCoV-19/England/CAMC-A80CFB/2020|EPI_ISL_610666|20 ···················· ···················· ····················· ···················· ···················· ····················
O 4510 hCoV-19/England/CAMC-A82E3F/2020|EPI_ISL_610667|20 ···················· ·A·················· ····················· ···················· ···················· ···A················
O 4511 hCoV-19/England/CAMC-A81B1B/2020|EPI_ISL_610668|20 ···················· ···················· ····················· ···················· ···················· ····················
O 4512 hCoV-19/England/CAMC-A82E6C/2020|EPI_ISL_610669|20 ···················· ···················· ····················· ···················· ···················· ····················
O 4513 hCoV-19/England/CAMC-A82FF0/2020|EPI_ISL_610670|20 ···················· ···················· ····················· ···················· ···················· ····················
O 4514 hCoV-19/England/CAMC-A80C91/2020|EPI_ISL_610671|20 ···················· ···················· ····················· ···················· ···················· ····················
O 4515 hCoV-19/England/CAMC-A82F87/2020|EPI_ISL_610672|20 ···················· ···················· ····················· ···················· ···················· ····················
O 4516 hCoV-19/England/CAMC-A82725/2020|EPI_ISL_610673|20 ···················· ·A·················· ····················· ···················· ···················· ····················
O 4517 hCoV-19/England/CAMC-A80FC5/2020|EPI_ISL_610674|20 ···················· ···················· ····················· ···················· ···················· ····················
O 4518 hCoV-19/England/CAMC-A81BEE/2020|EPI_ISL_610676|20 ···················· ·A·················· ····················· ···················· ···················· ····················
O 4519 hCoV-19/England/CAMC-A82CBD/2020|EPI_ISL_610677|20 ···················· ···················· ····················· ···················· ···················· ····················
O 4520 hCoV-19/England/CAMC-A82497/2020|EPI_ISL_610700|20 ···················· ···················· ····················· ···················· ···················· ····················
O 4521 hCoV-19/England/CAMC-A8227F/2020|EPI_ISL_610701|20 ···················· ···················· ····················· ···················· ···················· ····················
O 4522 hCoV-19/England/CAMC-A81CEB/2020|EPI_ISL_610703|20 ···················· ···················· ····················· ···················· ···················· ····················
O 4523 hCoV-19/England/CAMC-A822BB/2020|EPI_ISL_610704|20 ···················· ···················· ····················· ···················· ···················· ····················
O 4524 hCoV-19/England/CAMC-A80F98/2020|EPI_ISL_610705|20 ···················· ···················· ····················· ···················· ···················· ····················
O 4525 hCoV-19/England/CAMC-A82AFF/2020|EPI_ISL_610706|20 ···················· ···················· ····················· ···················· ···················· ····················
O 4526 hCoV-19/England/CAMC-A82488/2020|EPI_ISL_610707|20 ···················· ···················· ····················· ···················· ···················· ····················
O 4527 hCoV-19/England/CAMC-A82E7B/2020|EPI_ISL_610708|20 ···················· ···················· ····················· ···················· ···················· ····················
O 4528 hCoV-19/England/CAMC-A81AE1/2020|EPI_ISL_610710|20 ···················· ···················· ····················· ···················· ···················· ····················
O 4529 hCoV-19/England/CAMB-1B6D4D/2020|EPI_ISL_611965|20 ···················· ···················· ····················· ···················· ···················· ····················
O 4530 hCoV-19/England/CAMB-1B6D6B/2020|EPI_ISL_611966|20 ···················· ···················· ····················· ···················· ···················· ····················
O 4531 hCoV-19/England/CAMB-1B6D7A/2020|EPI_ISL_611967|20 ···················· ···················· ····················· ···················· ···················· ····················
O 4532 hCoV-19/England/MILK-A19538/2020|EPI_ISL_611968|20 ···················· ···················· ····················· ···················· ···················· ····················
O 4533 hCoV-19/England/MILK-A18D22/2020|EPI_ISL_611969|20 ···················· ·A·················· ····················· ···················· ···················· ···A················
O 4534 hCoV-19/England/MILK-A18DAA/2020|EPI_ISL_611970|20 ···················· ·A·················· ····················· ···················· ···················· ···A················
O 4535 hCoV-19/England/PORT-2D466B/2020|EPI_ISL_611971|20 ···················· ·A·················· ····················· ···················· ···················· ···A················
O 4536 hCoV-19/Wales/PHWC-47F563/2020|EPI_ISL_611972|2020 ···················· ···················· ····················· ···················· ···················· ····················
O 4537 hCoV-19/Wales/PHWC-481328/2020|EPI_ISL_611973|2020 ···················· ···················· ····················· ···················· ···················· ····················
O 4538 hCoV-19/USA/MI-MDHHS-SC22169/2020|EPI_ISL_614220|2 ···················· ·············A······ ····················· ···················· ···················· ····················
O 4539 hCoV-19/USA/MI-MDHHS-SC22170/2020|EPI_ISL_614221|2 ···················· ···················· ····················· ···················· ···················· ····················
O 4540 hCoV-19/USA/MI-MDHHS-SC22171/2020|EPI_ISL_614222|2 ···················· ·A·················· ····················· ···················· ···················· ····················
O 4541 hCoV-19/USA/MI-MDHHS-SC22172/2020|EPI_ISL_614223|2 ···················· ···················· ····················· ···················· ···A················ ····················
O 4542 hCoV-19/USA/MI-MDHHS-SC22173/2020|EPI_ISL_614224|2 ···················· ···················· ····················· ···················· ···················· ····················
O 4543 hCoV-19/USA/MI-MDHHS-SC22174/2020|EPI_ISL_614225|2 ···················· ···················· ····················· ···················· ···················· ····················

| | | | | | | | | |
|---|---|---|---|---|---|---|---|---|
| O | 4544 | hCoV-19/USA/MI-MDHHS-SC22176/2020\|EPI_ISL_614227\|2 | ···················· | ···················· | ···················· | ···················· | ···················· | ···················· |
| O | 4545 | hCoV-19/USA/MI-MDHHS-SC22177/2020\|EPI_ISL_614228\|2 | ···················· | ···················· | ···················· | ···················· | ···················· | ···················· |
| O | 4546 | hCoV-19/USA/MI-MDHHS-SC22178/2020\|EPI_ISL_614229\|2 | ···················· | ···················· | ···················· | ···················· | ···················· | ···················· |
| O | 4547 | hCoV-19/USA/MI-MDHHS-SC22179/2020\|EPI_ISL_614230\|2 | ···················· | ···················· | ···················· | ···················· | ···················· | ···················· |
| O | 4548 | hCoV-19/USA/MI-MDHHS-SC22180/2020\|EPI_ISL_614231\|2 | ···················· | ···················· | ···················· | ···················· | ···················· | ···················· |
| O | 4549 | hCoV-19/USA/MI-MDHHS-SC22181/2020\|EPI_ISL_614232\|2 | ···················· | ·A·················· | ···················· | ···················· | ···················· | ···················· |
| O | 4550 | hCoV-19/USA/MI-MDHHS-SC22182/2020\|EPI_ISL_614233\|2 | ···················· | ···················· | ···················· | ···················· | ···················· | ···················· |
| O | 4551 | hCoV-19/USA/MI-MDHHS-SC22183/2020\|EPI_ISL_614234\|2 | ···················· | ···················· | ···················· | ···················· | ···················· | ···················· |
| O | 4552 | hCoV-19/USA/MI-MDHHS-SC22184/2020\|EPI_ISL_614235\|2 | ···················· | ···················· | ···················· | ···················· | ···················· | ···················· |
| O | 4553 | hCoV-19/USA/MI-MDHHS-SC22185/2020\|EPI_ISL_614236\|2 | ···················· | ···················· | ···················· | ···················· | ···················· | ···················· |
| O | 4554 | hCoV-19/USA/MI-MDHHS-SC22186/2020\|EPI_ISL_614237\|2 | ···················· | ···················· | ···················· | ···················· | ···················· | ···················· |
| O | 4555 | hCoV-19/USA/MI-MDHHS-SC22187/2020\|EPI_ISL_614238\|2 | ···················· | ···················· | ···················· | ···················· | ···················· | ···················· |
| O | 4556 | hCoV-19/USA/MI-MDHHS-SC22188/2020\|EPI_ISL_614239\|2 | ···················· | ···················· | ···················· | ···················· | ···················· | ···A················ |
| O | 4557 | hCoV-19/USA/MI-MDHHS-SC22189/2020\|EPI_ISL_614240\|2 | ···················· | ·A·················· | ···················· | ···················· | ···················· | ···················· |
| O | 4558 | hCoV-19/USA/MI-MDHHS-SC22190/2020\|EPI_ISL_614241\|2 | ···················· | ·············A······ | ···················· | ···················· | ···················· | ···················· |
| O | 4559 | hCoV-19/USA/MI-MDHHS-SC22191/2020\|EPI_ISL_614242\|2 | ···················· | ···················· | ···················· | ···················· | ···················· | ···················· |
| O | 4560 | hCoV-19/USA/MI-MDHHS-SC22192/2020\|EPI_ISL_614243\|2 | ···················· | ···················· | ···················· | ···················· | ···················· | ···················· |
| O | 4561 | hCoV-19/USA/MI-MDHHS-SC22193/2020\|EPI_ISL_614244\|2 | ···················· | ···················· | ···················· | ···················· | ···················· | ···················· |
| O | 4562 | hCoV-19/USA/MI-MDHHS-SC22194/2020\|EPI_ISL_614245\|2 | ···················· | ···················· | ···················· | ···················· | ···················· | ···················· |
| O | 4563 | hCoV-19/USA/MI-MDHHS-SC22195/2020\|EPI_ISL_614246\|2 | ···················· | ···················· | ···················· | ···················· | ···················· | ···················· |
| O | 4564 | hCoV-19/USA/WY-UNM-00129/2020\|EPI_ISL_614249\|2020- | ···················· | ···················· | ···················· | ···················· | ···················· | ···················· |
| O | 4565 | hCoV-19/USA/WY-UNM-00132/2020\|EPI_ISL_614250\|2020- | ···················· | ···················· | ···················· | ···················· | ···················· | ···················· |
| O | 4566 | hCoV-19/USA/WY-UNM-00133/2020\|EPI_ISL_614251\|2020- | ···················· | ···················· | ···················· | ···················· | ···················· | ···················· |
| O | 4567 | hCoV-19/USA/WY-UNM-00134/2020\|EPI_ISL_614252\|2020- | ···················· | ···················· | ···················· | ···················· | ···················· | ···················· |
| O | 4568 | hCoV-19/USA/WY-UNM-00130/2020\|EPI_ISL_614253\|2020- | ···················· | ···················· | ···················· | ···················· | ···················· | ···················· |
| O | 4569 | hCoV-19/USA/WY-UNM-00131/2020\|EPI_ISL_614254\|2020- | ···················· | ···················· | ···················· | ···················· | ···················· | ···················· |
| O | 4570 | hCoV-19/USA/WY-UNM-00135/2020\|EPI_ISL_614255\|2020- | ···················· | ···················· | ···················· | ···················· | ···················· | ···················· |
| O | 4571 | hCoV-19/USA/WY-UNM-00136/2020\|EPI_ISL_614256\|2020- | ···················· | ···················· | ···················· | ···················· | ···················· | ···················· |
| O | 4572 | hCoV-19/USA/WY-UNM-00137/2020\|EPI_ISL_614257\|2020- | ···················· | ···················· | ···················· | ···················· | ···················· | ···················· |
| O | 4573 | hCoV-19/USA/WY-UNM-00138/2020\|EPI_ISL_614258\|2020- | ···················· | ···················· | ···················· | ···················· | ···················· | ···················· |
| O | 4574 | hCoV-19/USA/WY-UNM-00140/2020\|EPI_ISL_614259\|2020- | ···················· | ···················· | ···················· | ···················· | ···················· | ···················· |
| O | 4575 | hCoV-19/USA/WY-UNM-00141/2020\|EPI_ISL_614260\|2020- | ···················· | ···················· | ···················· | ···················· | ···················· | ···················· |
| O | 4576 | hCoV-19/USA/WY-UNM-00142/2020\|EPI_ISL_614261\|2020- | ···················· | ···················· | ···················· | ···················· | ···················· | ···················· |
| O | 4577 | hCoV-19/USA/WY-UNM-00143/2020\|EPI_ISL_614262\|2020- | ···················· | ···················· | ···················· | ···················· | ···················· | ···················· |
| O | 4578 | hCoV-19/USA/WY-UNM-00144/2020\|EPI_ISL_614263\|2020- | ···················· | ···················· | ···················· | ···················· | ···················· | ···················· |
| O | 4579 | hCoV-19/USA/WY-UNM-00145/2020\|EPI_ISL_614264\|2020- | ···················· | ···················· | ···················· | ···················· | ···················· | ···················· |
| O | 4580 | hCoV-19/France/BRE-9325/2020\|EPI_ISL_614265\|2020-0 | ···················· | ···················· | ···················· | ···················· | ···················· | ···················· |
| O | 4581 | hCoV-19/France/BRE-9326/2020\|EPI_ISL_614266\|2020-0 | ···················· | ···················· | ···················· | ···················· | ···················· | ···················· |
| O | 4582 | hCoV-19/France/BRE-9327/2020\|EPI_ISL_614267\|2020-0 | ···················· | ···················· | ···················· | ···················· | ···················· | ···················· |
| O | 4583 | hCoV-19/France/BRE-9329/2020\|EPI_ISL_614268\|2020-0 | ···················· | ···················· | ···················· | ···················· | ···················· | ···················· |
| O | 4584 | hCoV-19/France/BRE-9330/2020\|EPI_ISL_614269\|2020-0 | ···················· | ···················· | ···················· | ···················· | ···················· | ···················· |
| O | 4585 | hCoV-19/France/BRE-9331/2020\|EPI_ISL_614270\|2020-0 | ···················· | ···················· | ···················· | ···················· | ···················· | ···················· |
| O | 4586 | hCoV-19/France/BRE-9332/2020\|EPI_ISL_614271\|2020-0 | ···················· | ···················· | ···················· | ···················· | ···················· | ···················· |
| O | 4587 | hCoV-19/France/BRE-9333/2020\|EPI_ISL_614272\|2020-0 | ···················· | ···················· | ···················· | ···················· | ···················· | ···················· |
| O | 4588 | hCoV-19/France/BRE-9335/2020\|EPI_ISL_614273\|2020-0 | ···················· | ···················· | ···················· | ···················· | ···················· | ···················· |
| O | 4589 | hCoV-19/France/BRE-9339/2020\|EPI_ISL_614274\|2020-0 | ···················· | ···················· | ···················· | ···················· | ···················· | ···················· |
| O | 4590 | hCoV-19/France/BRE-9343/2020\|EPI_ISL_614275\|2020-0 | ···················· | ···················· | ···················· | ···················· | ···················· | ···················· |
| O | 4591 | hCoV-19/France/BRE-9344/2020\|EPI_ISL_614276\|2020-0 | ···················· | ···················· | ···················· | ···················· | ···················· | ···················· |
| O | 4592 | hCoV-19/France/BRE-9346/2020\|EPI_ISL_614277\|2020-0 | ···················· | ···················· | ···················· | ···················· | ···················· | ···················· |
| O | 4593 | hCoV-19/France/BRE-9348/2020\|EPI_ISL_614278\|2020-0 | ···················· | ···················· | ···················· | ···················· | ···················· | ···················· |
| O | 4594 | hCoV-19/France/BRE-9351/2020\|EPI_ISL_614279\|2020-0 | ···················· | ···················· | ···················· | ···················· | ···················· | ···················· |
| O | 4595 | hCoV-19/France/BRE-9353/2020\|EPI_ISL_614280\|2020-0 | ···················· | ···················· | ···················· | ···················· | ···················· | ···················· |
| O | 4596 | hCoV-19/France/BRE-9376/2020\|EPI_ISL_614281\|2020-0 | ···················· | ···················· | ···················· | ···················· | ···················· | ···················· |
| O | 4597 | hCoV-19/France/GES-9388/2020\|EPI_ISL_614282\|2020-1 | ···················· | ···················· | ···················· | ···················· | ···················· | ···················· |
| O | 4598 | hCoV-19/France/BFC-9406/2020\|EPI_ISL_614283\|2020-1 | ···················· | ···················· | ···················· | ···················· | ···················· | ···················· |
| O | 4599 | hCoV-19/France/HDF-9440/2020\|EPI_ISL_614284\|2020-1 | ···················· | ···················· | ···················· | ···················· | ···················· | ···················· |
| O | 4600 | hCoV-19/France/IDF-9449/2020\|EPI_ISL_614285\|2020-1 | ···················· | ···················· | ···················· | ···················· | ···················· | ···················· |
| O | 4601 | hCoV-19/France/CVL-9458/2020\|EPI_ISL_614286\|2020-1 | ···················· | ···················· | ···················· | ···················· | ···················· | ···················· |
| O | 4602 | hCoV-19/France/IDF-9462/2020\|EPI_ISL_614287\|2020-1 | ···················· | ···················· | ···················· | ···················· | ···················· | ···················· |
| O | 4603 | hCoV-19/France/CVL-9469/2020\|EPI_ISL_614288\|2020-1 | ···················· | ···················· | ···················· | ···················· | ···················· | ···················· |
| O | 4604 | hCoV-19/France/CVL-9471/2020\|EPI_ISL_614289\|2020-1 | ···················· | ···················· | ···················· | ···················· | ···················· | ···················· |
| O | 4605 | hCoV-19/France/CVL-9474/2020\|EPI_ISL_614290\|2020-1 | ···················· | ···················· | ···················· | ···················· | ···················· | ···················· |
| O | 4606 | hCoV-19/France/IDF-9475/2020\|EPI_ISL_614291\|2020-1 | ···················· | ···················· | ···················· | ···················· | ···················· | ···················· |
| O | 4607 | hCoV-19/France/GES-9488/2020\|EPI_ISL_614292\|2020-1 | ···················· | ···················· | ···················· | ···················· | ···················· | ···················· |
| O | 4608 | hCoV-19/France/IDF-9492/2020\|EPI_ISL_614293\|2020-1 | ···················· | ···················· | ···················· | ···················· | ···················· | ···················· |
| O | 4609 | hCoV-19/England/ALDP-A6F8D0/2020\|EPI_ISL_607022\|20 | ···················· | ···················· | ···················· | ···················· | ···················· | ···················· |
| O | 4610 | hCoV-19/England/ALDP-A6E5DA/2020\|EPI_ISL_607030\|20 | ···················· | ···················· | ···················· | ···················· | ···················· | ···················· |
| O | 4611 | hCoV-19/England/ALDP-A6FBAA/2020\|EPI_ISL_607031\|20 | ···················· | ···················· | ···················· | ···················· | ···················· | ···················· |
| O | 4612 | hCoV-19/England/ALDP-A6DD97/2020\|EPI_ISL_607032\|20 | ···················· | ···················· | ···················· | ···················· | ···················· | ···················· |
| O | 4613 | hCoV-19/England/ALDP-A6FC4D/2020\|EPI_ISL_607033\|20 | ···················· | ···················· | ···················· | ···················· | ···················· | ···················· |
| O | 4614 | hCoV-19/England/ALDP-A6E4EC/2020\|EPI_ISL_607034\|20 | ···················· | ···················· | ···················· | ···················· | ···················· | ···················· |
| O | 4615 | hCoV-19/England/ALDP-A6CC5F/2020\|EPI_ISL_607035\|20 | ···················· | ···················· | ···················· | ···················· | ···················· | ···················· |
| O | 4616 | hCoV-19/England/ALDP-A6D957/2020\|EPI_ISL_607036\|20 | ···················· | ···················· | ···················· | ···················· | ···················· | ···················· |
| O | 4617 | hCoV-19/England/ALDP-A6EAEA/2020\|EPI_ISL_607037\|20 | ···················· | ···················· | ···················· | ···················· | ···················· | ···················· |
| O | 4618 | hCoV-19/England/ALDP-A6EDA5/2020\|EPI_ISL_607038\|20 | ···················· | ···················· | ···················· | ···················· | ···················· | ···················· |
| O | 4619 | hCoV-19/England/ALDP-A6D5DB/2020\|EPI_ISL_607039\|20 | ···················· | ···················· | ···················· | ···················· | ···················· | ···················· |
| O | 4620 | hCoV-19/England/ALDP-A6FD86/2020\|EPI_ISL_607040\|20 | ···················· | ···················· | ···················· | ···················· | ···················· | ···················· |
| O | 4621 | hCoV-19/England/ALDP-A6EEA2/2020\|EPI_ISL_607041\|20 | ···················· | ···················· | ···················· | ···················· | ···················· | ···················· |
| O | 4622 | hCoV-19/England/ALDP-A6D614/2020\|EPI_ISL_607042\|20 | ···················· | ···················· | ···················· | ···················· | ···················· | ···················· |
| O | 4623 | hCoV-19/England/ALDP-A6E31C/2020\|EPI_ISL_607043\|20 | ···················· | ···················· | ···················· | ···················· | ···················· | ···················· |

| | | | | | | | |
|---|---|---|---|---|---|---|---|
| O | 4624 hCoV-19/England/ALDP-A6C92B/2020\|EPI_ISL_607044\|20 | ···················· | ·A·················· | ···················· | ···················· | ···················· | ··A················· |
| O | 4625 hCoV-19/England/ALDP-A6D650/2020\|EPI_ISL_607045\|20 | ···················· | ···················· | ···················· | ···················· | ···················· | ···················· |
| O | 4626 hCoV-19/England/ALDP-A6E376/2020\|EPI_ISL_607046\|20 | ···················· | ···················· | ···················· | ···················· | ···················· | ···················· |
| O | 4627 hCoV-19/England/ALDP-A6EE84/2020\|EPI_ISL_607047\|20 | ···················· | ···················· | ···················· | ···················· | ···················· | ···················· |
| O | 4628 hCoV-19/England/ALDP-A6E59E/2020\|EPI_ISL_607048\|20 | ···················· | ···················· | ···················· | ···················· | ···················· | ···················· |
| O | 4629 hCoV-19/England/ALDP-A6CC7D/2020\|EPI_ISL_607049\|20 | ···················· | ···················· | ···················· | ···················· | ···················· | ···················· |
| O | 4630 hCoV-19/England/ALDP-A6F876/2020\|EPI_ISL_607050\|20 | ···················· | ···················· | ···················· | ···················· | ···················· | ···················· |
| O | 4631 hCoV-19/England/ALDP-A6FB5F/2020\|EPI_ISL_607051\|20 | ···················· | ···················· | ···················· | ···················· | ···················· | ···················· |
| O | 4632 hCoV-19/England/ALDP-A6EBC9/2020\|EPI_ISL_607052\|20 | ···················· | ···················· | ···················· | ···················· | ···················· | ···················· |
| O | 4633 hCoV-19/England/ALDP-A6DC6D/2020\|EPI_ISL_607053\|20 | ···················· | ···················· | ···················· | ···················· | ···················· | ···················· |
| O | 4634 hCoV-19/England/ALDP-A6E5BC/2020\|EPI_ISL_607054\|20 | ···················· | ···················· | ···················· | ···················· | ···················· | ···················· |
| O | 4635 hCoV-19/England/ALDP-A6EDC3/2020\|EPI_ISL_607055\|20 | ···················· | ···················· | ···················· | ···················· | ···················· | ···················· |
| O | 4636 hCoV-19/England/ALDP-A6FA8F/2020\|EPI_ISL_607056\|20 | ···················· | ···················· | ···················· | ···················· | ···················· | ···················· |
| O | 4637 hCoV-19/England/ALDP-A6F71F/2020\|EPI_ISL_607057\|20 | ···················· | ···················· | ···················· | ···················· | ···················· | ···················· |
| O | 4638 hCoV-19/England/ALDP-A6EE75/2020\|EPI_ISL_607058\|20 | ···················· | ···················· | ···················· | ···················· | ···················· | ···················· |
| O | 4639 hCoV-19/England/ALDP-A6EBF6/2020\|EPI_ISL_607059\|20 | ···················· | ·A·················· | ···················· | ···················· | ···················· | ···················· |
| O | 4640 hCoV-19/England/ALDP-A6FB31/2020\|EPI_ISL_607060\|20 | ···················· | ···················· | ···················· | ···················· | ···················· | ···················· |
| O | 4641 hCoV-19/England/ALDP-A6E5E9/2020\|EPI_ISL_607061\|20 | ···················· | ···················· | ···················· | ···················· | ···················· | ···················· |
| O | 4642 hCoV-19/England/ALDP-A6F73D/2020\|EPI_ISL_607062\|20 | ···················· | ···················· | ···················· | ···················· | ···················· | ···················· |
| O | 4643 hCoV-19/England/ALDP-A6D9C0/2020\|EPI_ISL_607063\|20 | ···················· | ···················· | ···················· | ···················· | ···················· | ···················· |
| O | 4644 hCoV-19/England/ALDP-A6F919/2020\|EPI_ISL_607064\|20 | ···················· | ···················· | ···················· | ···················· | ···················· | ···················· |
| O | 4645 hCoV-19/England/ALDP-A6C703/2020\|EPI_ISL_607065\|20 | ···················· | ···················· | ···················· | ···················· | ···················· | ···················· |
| O | 4646 hCoV-19/England/ALDP-A6E112/2020\|EPI_ISL_607066\|20 | ···················· | ···················· | ···················· | ···················· | ···················· | ···················· |
| O | 4647 hCoV-19/England/ALDP-A6FD0E/2020\|EPI_ISL_607067\|20 | ···················· | ···················· | ···················· | ···················· | ···················· | ···················· |
| O | 4648 hCoV-19/England/ALDP-A6DA09/2020\|EPI_ISL_607068\|20 | ···················· | ···················· | ···················· | ···················· | ···················· | ···················· |
| O | 4649 hCoV-19/England/ALDP-A6CA46/2020\|EPI_ISL_607069\|20 | ···················· | ···················· | ···················· | ···················· | ···················· | ···················· |
| O | 4650 hCoV-19/England/ALDP-A6D66F/2020\|EPI_ISL_607070\|20 | ···················· | ···················· | ···················· | ···················· | ···················· | ···················· |
| O | 4651 hCoV-19/England/ALDP-A6F278/2020\|EPI_ISL_607071\|20 | ···················· | ···················· | ···················· | ···················· | ···················· | ···················· |
| O | 4652 hCoV-19/England/ALDP-A6E631/2020\|EPI_ISL_607072\|20 | ···················· | ···················· | ···················· | ···················· | ···················· | ···················· |
| O | 4653 hCoV-19/England/ALDP-A6C9B2/2020\|EPI_ISL_607073\|20 | ···················· | ···················· | ···················· | ···················· | ···················· | ···················· |
| O | 4654 hCoV-19/England/ALDP-A6DDC4/2020\|EPI_ISL_607074\|20 | ···················· | ···················· | ···················· | ···················· | ···················· | ···················· |
| O | 4655 hCoV-19/England/ALDP-A6FB6E/2020\|EPI_ISL_607075\|20 | ···················· | ···················· | ···················· | ···················· | ···················· | ···················· |
| O | 4656 hCoV-19/England/ALDP-A6DCD6/2020\|EPI_ISL_607076\|20 | ···················· | ···················· | ···················· | ···················· | ···················· | ···················· |
| O | 4657 hCoV-19/England/ALDP-A6DC9A/2020\|EPI_ISL_607077\|20 | ···················· | ···················· | ···················· | ···················· | ···················· | ···················· |
| O | 4658 hCoV-19/England/ALDP-A6F8B2/2020\|EPI_ISL_607078\|20 | ···················· | ···················· | ···················· | ···················· | ···················· | ···················· |
| O | 4659 hCoV-19/England/ALDP-A6F384/2020\|EPI_ISL_607079\|20 | ···················· | ···················· | ···················· | ···················· | ···················· | ···················· |
| O | 4660 hCoV-19/England/ALDP-A6CBF8/2020\|EPI_ISL_607080\|20 | ···················· | ···················· | ···················· | ···················· | ···················· | ···················· |
| O | 4661 hCoV-19/England/ALDP-A6FCE3/2020\|EPI_ISL_607082\|20 | ···················· | ···················· | ···················· | ···················· | ···················· | ···················· |
| O | 4662 hCoV-19/England/ALDP-A6EE93/2020\|EPI_ISL_607083\|20 | ···················· | ···················· | ···················· | ···················· | ···················· | ···················· |
| O | 4663 hCoV-19/England/ALDP-A6EEDF/2020\|EPI_ISL_607084\|20 | ···················· | ···················· | ···················· | ···················· | ···················· | ···················· |
| O | 4664 hCoV-19/England/ALDP-A6EF09/2020\|EPI_ISL_607085\|20 | ···················· | ···················· | ···················· | ···················· | ···················· | ···················· |
| O | 4665 hCoV-19/England/ALDP-A6F894/2020\|EPI_ISL_607086\|20 | ···················· | ···················· | ···················· | ···················· | ···················· | ···················· |
| O | 4666 hCoV-19/England/ALDP-A6E24C/2020\|EPI_ISL_607087\|20 | ···················· | ···················· | ···················· | ···················· | ···················· | ···················· |
| O | 4667 hCoV-19/England/ALDP-A6CC40/2020\|EPI_ISL_607088\|20 | ···················· | ···················· | ···················· | ···················· | ···················· | ···················· |
| O | 4668 hCoV-19/England/ALDP-A6EB8D/2020\|EPI_ISL_607089\|20 | ···················· | ···················· | ···················· | ···················· | ···················· | ···················· |
| O | 4669 hCoV-19/England/ALDP-A6FC89/2020\|EPI_ISL_607090\|20 | ···················· | ···················· | ···················· | ···················· | ···················· | ···················· |
| O | 4670 hCoV-19/England/ALDP-A6FDA4/2020\|EPI_ISL_607091\|20 | ···················· | ···················· | ···················· | ···················· | ···················· | ···················· |
| O | 4671 hCoV-19/England/ALDP-A6F90A/2020\|EPI_ISL_607092\|20 | ···················· | ···················· | ···················· | ···················· | ···················· | ···················· |
| O | 4672 hCoV-19/England/ALDP-A6FD68/2020\|EPI_ISL_607093\|20 | ···················· | ···················· | ···················· | ···················· | ···················· | ···················· |
| O | 4673 hCoV-19/England/ALDP-A6FCB6/2020\|EPI_ISL_607094\|20 | ···················· | ···················· | ···················· | ···················· | ···················· | ···················· |
| O | 4674 hCoV-19/England/ALDP-A6F1D5/2020\|EPI_ISL_607096\|20 | ···················· | ···················· | ···················· | ···················· | ···················· | ···················· |
| O | 4675 hCoV-19/England/ALDP-A6F31B/2020\|EPI_ISL_607097\|20 | ···················· | ···················· | ···················· | ···················· | ···················· | ···················· |
| O | 4676 hCoV-19/England/ALDP-A6EDB4/2020\|EPI_ISL_607098\|20 | ···················· | ···················· | ···················· | ···················· | ···················· | ···················· |
| O | 4677 hCoV-19/England/ALDP-A6F199/2020\|EPI_ISL_607099\|20 | ···················· | ···················· | ···················· | ···················· | ···················· | ···················· |
| O | 4678 hCoV-19/England/ALDP-A6E84A/2020\|EPI_ISL_607801\|20 | ···················· | ···················· | ···················· | ···················· | ···················· | ···················· |
| O | 4679 hCoV-19/England/ALDP-A70292/2020\|EPI_ISL_607802\|20 | ···················· | ···················· | ···················· | ···················· | ···················· | ···················· |
| O | 4680 hCoV-19/England/ALDP-A6F9A0/2020\|EPI_ISL_607803\|20 | ···················· | ···················· | ···················· | ···················· | ···················· | ···················· |
| O | 4681 hCoV-19/England/ALDP-A6F0F6/2020\|EPI_ISL_607804\|20 | ···················· | ···················· | ···················· | ···················· | ···················· | ···················· |
| O | 4682 hCoV-19/England/ALDP-A6E09D/2020\|EPI_ISL_607805\|20 | ···················· | ···················· | ···················· | ···················· | ···················· | ···················· |
| O | 4683 hCoV-19/England/ALDP-A6D544/2020\|EPI_ISL_608300\|20 | ···················· | ···················· | ···················· | ···················· | ···················· | ···················· |
| O | 4684 hCoV-19/England/ALDP-A6FE92/2020\|EPI_ISL_608301\|20 | ···················· | ···················· | ···················· | ···················· | ···················· | ···················· |
| O | 4685 hCoV-19/England/ALDP-A6CE95/2020\|EPI_ISL_608302\|20 | ···················· | ···················· | ···················· | ···················· | ···················· | ···················· |
| O | 4686 hCoV-19/England/ALDP-A6D4A1/2020\|EPI_ISL_608303\|20 | ···················· | ···················· | ···················· | ···················· | ···················· | ···················· |
| O | 4687 hCoV-19/England/ALDP-A6CF38/2020\|EPI_ISL_608304\|20 | ···················· | ···················· | ···················· | ···················· | ···················· | ···················· |
| O | 4688 hCoV-19/England/ALDP-A70001/2020\|EPI_ISL_608305\|20 | ···················· | ···················· | ···················· | ···················· | ···················· | ···················· |
| O | 4689 hCoV-19/England/ALDP-A6D80F/2020\|EPI_ISL_608306\|20 | ···················· | ···················· | ···················· | ···················· | ······T············· | ···················· |
| O | 4690 hCoV-19/England/ALDP-A6CFB0/2020\|EPI_ISL_608307\|20 | ···················· | ···················· | ···················· | ···················· | ···················· | ···················· |
| O | 4691 hCoV-19/England/ALDP-A6FE47/2020\|EPI_ISL_608308\|20 | ···················· | ···················· | ···················· | ···················· | ···················· | ···················· |
| O | 4692 hCoV-19/England/ALDP-A6CD89/2020\|EPI_ISL_608309\|20 | ···················· | ···················· | ···················· | ···················· | ···················· | ···················· |
| O | 4693 hCoV-19/England/ALDP-A70924/2020\|EPI_ISL_608311\|20 | ···················· | ···················· | ···················· | ···················· | ···················· | ···················· |
| O | 4694 hCoV-19/England/ALDP-A6D16E/2020\|EPI_ISL_608312\|20 | ···················· | ···················· | ···················· | ···················· | ···················· | ···················· |
| O | 4695 hCoV-19/England/ALDP-A6FFDB/2020\|EPI_ISL_608313\|20 | ···················· | ···················· | ···················· | ···················· | ···················· | ···················· |
| O | 4696 hCoV-19/England/ALDP-A6D7C6/2020\|EPI_ISL_608314\|20 | ···················· | ···················· | ···················· | ···················· | ···················· | ···················· |
| O | 4697 hCoV-19/England/ALDP-A6D17D/2020\|EPI_ISL_608315\|20 | ···················· | ···················· | ···················· | ···················· | ···················· | ···················· |
| O | 4698 hCoV-19/England/ALDP-A6D1F5/2020\|EPI_ISL_608316\|20 | ···················· | ···················· | ···················· | ···················· | ···················· | ···················· |
| O | 4699 hCoV-19/England/ALDP-A6C3F0/2020\|EPI_ISL_608317\|20 | ···················· | ···················· | ···················· | ···················· | ···················· | ···················· |
| O | 4700 hCoV-19/England/ALDP-A6CD10/2020\|EPI_ISL_608318\|20 | ···················· | ···················· | ···················· | ···················· | ···················· | ··A················· |
| O | 4701 hCoV-19/England/ALDP-A6FE56/2020\|EPI_ISL_608319\|20 | ···················· | ···················· | ···················· | ···················· | ···················· | ···················· |
| O | 4702 hCoV-19/England/ALDP-A68B92/2020\|EPI_ISL_608361\|20 | ···················· | ···················· | ···················· | ···················· | ···················· | ···················· |
| O | 4703 hCoV-19/England/ALDP-A68F1E/2020\|EPI_ISL_608362\|20 | ···················· | ···················· | ···················· | ···················· | ···················· | ···················· |

| | | | | | | | | |
|---|---|---|---|---|---|---|---|---|
| O | 4704 | hCoV-19/England/ALDP-A68E11/2020\|EPI_ISL_608364\|20 | ···················· | ···················· | ····················· | ···················· | ···················· | ···················· |
| O | 4705 | hCoV-19/England/ALDP-A68804/2020\|EPI_ISL_608365\|20 | ···················· | ···················· | ····················· | ···················· | ···················· | ···················· |
| O | 4706 | hCoV-19/England/ALDP-A67F1F/2020\|EPI_ISL_608366\|20 | ···················· | ···················· | ····················· | ···················· | ···················· | ···················· |
| O | 4707 | hCoV-19/England/ALDP-A683A9/2020\|EPI_ISL_608367\|20 | ···················· | ···················· | ····················· | ···················· | ···················· | ···················· |
| O | 4708 | hCoV-19/England/CAMC-A65096/2020\|EPI_ISL_608368\|20 | ···················· | ···················· | ····················· | ···················· | ···················· | ···················· |
| O | 4709 | hCoV-19/England/ALDP-A68CBD/2020\|EPI_ISL_608369\|20 | ···················· | ···················· | ····················· | ···················· | ···················· | ···················· |
| O | 4710 | hCoV-19/England/CAMC-A64AE4/2020\|EPI_ISL_608370\|20 | ···················· | ···················· | ····················· | ···················· | ···················· | ···················· |
| O | 4711 | hCoV-19/England/ALDP-A687BC/2020\|EPI_ISL_608371\|20 | ···················· | ···················· | ····················· | ···················· | ···················· | ···················· |
| O | 4712 | hCoV-19/England/ALDP-A68479/2020\|EPI_ISL_608372\|20 | ···················· | ···················· | ····················· | ···················· | ···················· | ···················· |
| O | 4713 | hCoV-19/England/CAMC-A655A6/2020\|EPI_ISL_608373\|20 | ···················· | ·A·················· | ····················· | ···················· | ···················· | ···A················ |
| O | 4714 | hCoV-19/England/ALDP-A68F0F/2020\|EPI_ISL_608374\|20 | ···················· | ···················· | ····················· | ···················· | ···················· | ···················· |
| O | 4715 | hCoV-19/England/ALDP-A68E7B/2020\|EPI_ISL_608375\|20 | ···················· | ···················· | ····················· | ···················· | ···················· | ···················· |
| O | 4716 | hCoV-19/England/ALDP-A6888C/2020\|EPI_ISL_608376\|20 | ···················· | ···················· | ····················· | ···················· | ···················· | ···················· |
| O | 4717 | hCoV-19/England/ALDP-A68206/2020\|EPI_ISL_608377\|20 | ···················· | ···················· | ····················· | ···················· | ···················· | ···················· |
| O | 4718 | hCoV-19/England/ALDP-A685C1/2020\|EPI_ISL_608378\|20 | ···················· | ···················· | ····················· | ···················· | ···················· | ···················· |
| O | 4719 | hCoV-19/England/ALDP-A684A6/2020\|EPI_ISL_608380\|20 | ···················· | ···················· | ····················· | ···················· | ···················· | ···················· |
| O | 4720 | hCoV-19/England/CAMC-A65333/2020\|EPI_ISL_608381\|20 | ···················· | ···················· | ····················· | ···················· | ···················· | ···················· |
| O | 4721 | hCoV-19/England/CAMC-A64F7C/2020\|EPI_ISL_608382\|20 | ···················· | ···················· | ····················· | ···················· | ···················· | ···················· |
| O | 4722 | hCoV-19/England/ALDP-A6889B/2020\|EPI_ISL_608384\|20 | ···················· | ···················· | ····················· | ···················· | ···················· | ···················· |
| O | 4723 | hCoV-19/England/ALDP-A68F5A/2020\|EPI_ISL_608385\|20 | ···················· | ···················· | ····················· | ···················· | ···················· | ···················· |
| O | 4724 | hCoV-19/England/ALDP-A684E2/2020\|EPI_ISL_608386\|20 | ···················· | ···················· | ····················· | ···················· | ···················· | ···················· |
| O | 4725 | hCoV-19/England/ALDP-A68400/2020\|EPI_ISL_608387\|20 | ···················· | ···················· | ····················· | ···················· | ···················· | ···················· |
| O | 4726 | hCoV-19/England/CAMC-A655F1/2020\|EPI_ISL_608388\|20 | ···················· | ···················· | ····················· | ···················· | ···················· | ···················· |
| O | 4727 | hCoV-19/England/CAMC-A64F5E/2020\|EPI_ISL_608389\|20 | ···················· | ···················· | ····················· | ···················· | ···················· | ···················· |
| O | 4728 | hCoV-19/England/ALDP-A68594/2020\|EPI_ISL_608390\|20 | ···················· | ···················· | ····················· | ···················· | ···················· | ···················· |
| O | 4729 | hCoV-19/England/CAMC-A6510C/2020\|EPI_ISL_608391\|20 | ···················· | ···················· | ····················· | ···················· | ···················· | ···················· |
| O | 4730 | hCoV-19/England/CAMC-A64E51/2020\|EPI_ISL_608392\|20 | ···················· | ···················· | ····················· | ···················· | ···················· | ···················· |
| O | 4731 | hCoV-19/England/CAMC-A65579/2020\|EPI_ISL_608393\|20 | ···················· | ···················· | ····················· | ···················· | ···················· | ···················· |
| O | 4732 | hCoV-19/England/CAMC-A6512A/2020\|EPI_ISL_608394\|20 | ···················· | ·A·················· | ····················· | ···················· | ···················· | ···················· |
| O | 4733 | hCoV-19/England/CAMC-A64DDC/2020\|EPI_ISL_608395\|20 | ···················· | ·A·················· | ····················· | ···················· | ···················· | ···A················ |
| O | 4734 | hCoV-19/England/CAMC-A6570A/2020\|EPI_ISL_608396\|20 | ···················· | ···················· | ····················· | ···················· | ···················· | ···················· |
| O | 4735 | hCoV-19/England/CAMC-A64ECA/2020\|EPI_ISL_608397\|20 | ···················· | ···················· | ····················· | ···················· | ···················· | ···················· |
| O | 4736 | hCoV-19/England/CAMC-A655C4/2020\|EPI_ISL_608398\|20 | ···················· | ·A·······A·········· | ····················· | ···················· | ···················· | ···················· |
| O | 4737 | hCoV-19/England/ALDP-A68C35/2020\|EPI_ISL_608399\|20 | ···················· | ···················· | ····················· | ···················· | ···················· | ···················· |
| O | 4738 | hCoV-19/England/CAMC-A3E8F8/2020\|EPI_ISL_609610\|20 | ···················· | ···················· | ····················· | ···················· | ···················· | ···················· |
| O | 4739 | hCoV-19/England/CAMC-A3E755/2020\|EPI_ISL_609611\|20 | ···················· | ···················· | ····················· | ···················· | ···················· | ···················· |
| O | 4740 | hCoV-19/England/CAMC-A3E4D6/2020\|EPI_ISL_609612\|20 | ···················· | ···················· | ····················· | ···················· | ···················· | ···················· |
| O | 4741 | hCoV-19/England/CAMC-A3E97D/2020\|EPI_ISL_609613\|20 | ···················· | ···················· | ····················· | ···················· | ···················· | ···················· |
| O | 4742 | hCoV-19/England/CAMC-A3FBB2/2020\|EPI_ISL_609614\|20 | ···················· | ···················· | ····················· | ···················· | ···················· | ···················· |
| O | 4743 | hCoV-19/England/CAMC-A3E658/2020\|EPI_ISL_609615\|20 | ···················· | ···················· | ····················· | ···················· | ···················· | ···················· |
| O | 4744 | hCoV-19/England/CAMC-A3EAF2/2020\|EPI_ISL_609616\|20 | ···················· | ···················· | ····················· | ···················· | ···················· | ···················· |
| O | 4745 | hCoV-19/England/CAMC-A3E9AA/2020\|EPI_ISL_609617\|20 | ···················· | ···················· | ····················· | ···················· | ···················· | ···················· |
| O | 4746 | hCoV-19/England/CAMC-A3F156/2020\|EPI_ISL_609618\|20 | ···················· | ···················· | ····················· | ···················· | ···················· | ···················· |
| O | 4747 | hCoV-19/England/CAMC-A3EDBD/2020\|EPI_ISL_609619\|20 | ···················· | ···················· | ····················· | ···················· | ···················· | ···················· |
| O | 4748 | hCoV-19/England/CAMC-A3F271/2020\|EPI_ISL_609620\|20 | ···················· | ···················· | ····················· | ···················· | ···················· | ···················· |
| O | 4749 | hCoV-19/England/CAMC-A3F350/2020\|EPI_ISL_609621\|20 | ···················· | ···················· | ····················· | ···················· | ···················· | ···················· |
| O | 4750 | hCoV-19/England/CAMC-A3FB76/2020\|EPI_ISL_609622\|20 | ···················· | ···················· | ····················· | ···················· | ···················· | ···················· |
| O | 4751 | hCoV-19/England/CAMC-A3F217/2020\|EPI_ISL_609623\|20 | ···················· | ···················· | ····················· | ···················· | ···················· | ···················· |
| O | 4752 | hCoV-19/England/CAMC-A3E5D3/2020\|EPI_ISL_609625\|20 | ···················· | ···················· | ····················· | ···················· | ···················· | ···················· |
| O | 4753 | hCoV-19/England/CAMC-A3F1B0/2020\|EPI_ISL_609626\|20 | ···················· | ···················· | ····················· | ···················· | ···················· | ···················· |
| O | 4754 | hCoV-19/England/CAMC-A3F2EA/2020\|EPI_ISL_609627\|20 | ···················· | ···················· | ····················· | ···················· | ···················· | ···················· |
| O | 4755 | hCoV-19/England/CAMC-A3E6B2/2020\|EPI_ISL_609628\|20 | ···················· | ···················· | ····················· | ···················· | ···················· | ···················· |
| O | 4756 | hCoV-19/England/CAMC-A3F7FA/2020\|EPI_ISL_609629\|20 | ···················· | ···················· | ····················· | ···················· | ···················· | ···················· |
| O | 4757 | hCoV-19/England/CAMC-A3EAC5/2020\|EPI_ISL_609630\|20 | ···················· | ···················· | ····················· | ···················· | ···················· | ···················· |
| O | 4758 | hCoV-19/England/CAMC-A3E782/2020\|EPI_ISL_609631\|20 | ···················· | ···················· | ····················· | ···················· | ···················· | ···················· |
| O | 4759 | hCoV-19/England/CAMC-A3EB77/2020\|EPI_ISL_609632\|20 | ···················· | ···················· | ····················· | ···················· | ···················· | ···················· |
| O | 4760 | hCoV-19/England/CAMC-A3EB68/2020\|EPI_ISL_609633\|20 | ···················· | ···················· | ····················· | ···················· | ···················· | ···················· |
| O | 4761 | hCoV-19/England/CAMC-A3FB67/2020\|EPI_ISL_609634\|20 | ···················· | ···················· | ····················· | ···················· | ···················· | ···················· |
| O | 4762 | hCoV-19/England/CAMC-A3F8E8/2020\|EPI_ISL_609635\|20 | ···················· | ···················· | ····················· | ···················· | ···················· | ···················· |
| O | 4763 | hCoV-19/England/CAMC-A3FA88/2020\|EPI_ISL_609636\|20 | ···················· | ···················· | ····················· | ···················· | ···················· | ···················· |
| O | 4764 | hCoV-19/England/CAMC-A3E4F4/2020\|EPI_ISL_609637\|20 | ···················· | ···················· | ····················· | ···················· | ···················· | ···················· |
| O | 4765 | hCoV-19/England/CAMC-A3E7BF/2020\|EPI_ISL_609638\|20 | ···················· | ·A·················· | ····················· | ···················· | ···················· | ···················· |
| O | 4766 | hCoV-19/England/CAMC-A3E9F5/2020\|EPI_ISL_609639\|20 | ···················· | ···················· | ····················· | ···················· | ···················· | ···················· |
| O | 4767 | hCoV-19/England/CAMC-A3ED44/2020\|EPI_ISL_609640\|20 | ···················· | ···················· | ····················· | ···················· | ···················· | ···················· |
| O | 4768 | hCoV-19/England/CAMC-A3F2BD/2020\|EPI_ISL_609641\|20 | ···················· | ···················· | ····················· | ···················· | ···················· | ···················· |
| O | 4769 | hCoV-19/England/CAMC-A3F6C0/2020\|EPI_ISL_609642\|20 | ···················· | ···················· | ····················· | ···················· | ···················· | ···················· |
| O | 4770 | hCoV-19/England/CAMC-A3EE32/2020\|EPI_ISL_609644\|20 | ···················· | ···················· | ····················· | ···················· | ···················· | ···················· |
| O | 4771 | hCoV-19/England/CAMC-A3F411/2020\|EPI_ISL_609645\|20 | ···················· | ···················· | ····················· | ···················· | ···················· | ···················· |
| O | 4772 | hCoV-19/England/CAMC-A3F129/2020\|EPI_ISL_609646\|20 | ···················· | ···················· | ····················· | ···················· | ···················· | ···················· |
| O | 4773 | hCoV-19/England/CAMC-A3ECCF/2020\|EPI_ISL_609647\|20 | ···················· | ···················· | ····················· | ···················· | ···················· | ···················· |
| O | 4774 | hCoV-19/England/CAMC-A3F4E4/2020\|EPI_ISL_609648\|20 | ···················· | ···················· | ····················· | ···················· | ···················· | ···················· |
| O | 4775 | hCoV-19/England/CAMC-A3E728/2020\|EPI_ISL_609649\|20 | ···················· | ···················· | ····················· | ···················· | ···················· | ···················· |
| O | 4776 | hCoV-19/England/CAMC-A3F745/2020\|EPI_ISL_609690\|20 | ···················· | ···················· | ····················· | ···················· | ···················· | ···················· |
| O | 4777 | hCoV-19/England/CAMC-A3E649/2020\|EPI_ISL_609691\|20 | ···················· | ···················· | ····················· | ···················· | ···················· | ···················· |
| O | 4778 | hCoV-19/England/CAMC-A3FA3D/2020\|EPI_ISL_609693\|20 | ···················· | ···················· | ····················· | ···················· | ···················· | ···················· |
| O | 4779 | hCoV-19/England/CAMC-A3F0E0/2020\|EPI_ISL_609694\|20 | ···················· | ···················· | ····················· | ···················· | ···················· | ···················· |
| O | 4780 | hCoV-19/England/CAMC-A3E6EF/2020\|EPI_ISL_609695\|20 | ···················· | ···················· | ····················· | ···················· | ···················· | ···················· |
| O | 4781 | hCoV-19/England/CAMC-A3E44F/2020\|EPI_ISL_609696\|20 | ···················· | ···················· | ····················· | ···················· | ···················· | ···················· |
| O | 4782 | hCoV-19/England/CAMC-A3F5A5/2020\|EPI_ISL_609697\|20 | ···················· | ···················· | ····················· | ···················· | ···················· | ···················· |
| O | 4783 | hCoV-19/England/CAMC-A3F709/2020\|EPI_ISL_609698\|20 | ···················· | ···················· | ····················· | ···················· | ···················· | ···················· |

| | | | | | | | | |
|---|---|---|---|---|---|---|---|---|
| O | 4784 | hCoV-19/England/CAMC-A3F930/2020\|EPI_ISL_609699\|20 | ···················· | ···················· | ···················· | ···················· | ···················· | ···················· |
| O | 4785 | hCoV-19/England/ALDP-A84A66/2020\|EPI_ISL_610600\|20 | ···················· | ···················· | ···················· | ···················· | ···················· | ···················· |
| O | 4786 | hCoV-19/England/ALDP-A860CC/2020\|EPI_ISL_610601\|20 | ···················· | ···················· | ···················· | ···················· | ···················· | ···················· |
| O | 4787 | hCoV-19/England/ALDP-A84C06/2020\|EPI_ISL_610604\|20 | ···················· | ···················· | ···················· | ···················· | ···················· | ···················· |
| O | 4788 | hCoV-19/England/ALDP-A85BEA/2020\|EPI_ISL_610605\|20 | ···················· | ···················· | ···················· | ···················· | ···················· | ···················· |
| O | 4789 | hCoV-19/England/ALDP-A85A47/2020\|EPI_ISL_610608\|20 | ···················· | ···················· | ···················· | ···················· | ···················· | ···················· |
| O | 4790 | hCoV-19/England/ALDP-A84AC0/2020\|EPI_ISL_610609\|20 | ···················· | ···················· | ···················· | ···················· | ···················· | ···················· |
| O | 4791 | hCoV-19/England/ALDP-A85F2A/2020\|EPI_ISL_610610\|20 | ···················· | ···················· | ···················· | ···················· | ···················· | ···················· |
| O | 4792 | hCoV-19/England/ALDP-A85D6C/2020\|EPI_ISL_610612\|20 | ···················· | ···················· | ···················· | ···················· | ···················· | ···················· |
| O | 4793 | hCoV-19/England/ALDP-A85BBD/2020\|EPI_ISL_610613\|20 | ···················· | ···················· | ···················· | ···················· | ···················· | ···················· |
| O | 4794 | hCoV-19/England/ALDP-A84D5E/2020\|EPI_ISL_610614\|20 | ···················· | ·A·················· | ···················· | ···················· | ···················· | ···A················ |
| O | 4795 | hCoV-19/England/ALDP-A85661/2020\|EPI_ISL_610616\|20 | ···················· | ···················· | ···················· | ···················· | ···················· | ···················· |
| O | 4796 | hCoV-19/England/ALDP-A85DA8/2020\|EPI_ISL_610617\|20 | ···················· | ···················· | ···················· | ···················· | ···················· | ···················· |
| O | 4797 | hCoV-19/England/ALDP-A84C9D/2020\|EPI_ISL_610618\|20 | ···················· | ···················· | ···················· | ···················· | ···················· | ···················· |
| O | 4798 | hCoV-19/England/ALDP-A859FF/2020\|EPI_ISL_610619\|20 | ···················· | ···················· | ···················· | ···················· | ···················· | ···················· |
| O | 4799 | hCoV-19/England/ALDP-A84E1F/2020\|EPI_ISL_610620\|20 | ···················· | ···················· | ···················· | ···················· | ···················· | ···················· |
| O | 4800 | hCoV-19/England/ALDP-A8585C/2020\|EPI_ISL_610621\|20 | ···················· | ···················· | ···················· | ···················· | ···················· | ···················· |
| O | 4801 | hCoV-19/England/ALDP-A84F2B/2020\|EPI_ISL_610622\|20 | ···················· | ·····C·············· | ···················· | ···················· | ···················· | ···················· |
| O | 4802 | hCoV-19/England/ALDP-A85D4E/2020\|EPI_ISL_610624\|20 | ···················· | ···················· | ···················· | ···················· | ···················· | ···················· |
| O | 4803 | hCoV-19/England/ALDP-A84AB1/2020\|EPI_ISL_610625\|20 | ···················· | ···················· | ···················· | ···················· | ···················· | ···················· |
| O | 4804 | hCoV-19/England/ALDP-A84D6D/2020\|EPI_ISL_610626\|20 | ···················· | ···················· | ···················· | ···················· | ···················· | ···················· |
| O | 4805 | hCoV-19/England/ALDP-A85DB7/2020\|EPI_ISL_610627\|20 | ···················· | ···················· | ···················· | ···················· | ···················· | ···················· |
| O | 4806 | hCoV-19/England/ALDP-A85801/2020\|EPI_ISL_610628\|20 | ···················· | ···················· | ···················· | ···················· | ···················· | ···················· |
| O | 4807 | hCoV-19/Wales/CAMC-A80FB6/2020\|EPI_ISL_610629\|2020 | ···················· | ···················· | ···················· | ···················· | ···················· | ···················· |
| O | 4808 | hCoV-19/England/CAMC-A82D9C/2020\|EPI_ISL_610630\|20 | ···················· | ···················· | ···················· | ···················· | ···················· | ···················· |
| O | 4809 | hCoV-19/England/CAMC-A82F4B/2020\|EPI_ISL_610631\|20 | ···················· | ···················· | ···················· | ···················· | ······T············· | ···················· |
| O | 4810 | hCoV-19/England/CAMC-A82558/2020\|EPI_ISL_610632\|20 | ···················· | ···················· | ···················· | ···················· | ···················· | ···················· |
| O | 4811 | hCoV-19/England/CAMC-A81C36/2020\|EPI_ISL_610633\|20 | ···················· | ···················· | ···················· | ···················· | ···················· | ···················· |
| O | 4812 | hCoV-19/England/CAMC-A81CAF/2020\|EPI_ISL_610678\|20 | ···················· | ···················· | ···················· | ···················· | ···················· | ···················· |
| O | 4813 | hCoV-19/England/CAMC-A811BF/2020\|EPI_ISL_610679\|20 | ···················· | ···················· | ···················· | ···················· | ······T············· | ···················· |
| O | 4814 | hCoV-19/England/CAMC-A81C18/2020\|EPI_ISL_610681\|20 | ···················· | ···················· | ···················· | ···················· | ···················· | ···················· |
| O | 4815 | hCoV-19/England/CAMC-A82CF9/2020\|EPI_ISL_610683\|20 | ···················· | ···················· | ···················· | ···················· | ···················· | ···················· |
| O | 4816 | hCoV-19/England/CAMC-A81AC3/2020\|EPI_ISL_610684\|20 | ···················· | ···················· | ···················· | ···················· | ···················· | ···················· |
| O | 4817 | hCoV-19/England/CAMC-A83038/2020\|EPI_ISL_610685\|20 | ···················· | ···················· | ···················· | ···················· | ···················· | ···················· |
| O | 4818 | hCoV-19/England/CAMC-A80F5C/2020\|EPI_ISL_610687\|20 | ···················· | ···················· | ···················· | ···················· | ···················· | ···················· |
| O | 4819 | hCoV-19/England/CAMC-A82CDB/2020\|EPI_ISL_610688\|20 | ···················· | ···················· | ···················· | ···················· | ···················· | ···················· |
| O | 4820 | hCoV-19/England/CAMC-A81049/2020\|EPI_ISL_610689\|20 | ···················· | ·A·················· | ···················· | ···················· | ···················· | ···A················ |
| O | 4821 | hCoV-19/England/CAMC-A82EA8/2020\|EPI_ISL_610691\|20 | ···················· | ···················· | ···················· | ···················· | ···················· | ···················· |
| O | 4822 | hCoV-19/England/CAMC-A81FD3/2020\|EPI_ISL_610692\|20 | ···················· | ···················· | ···················· | ···················· | ···················· | ···················· |
| O | 4823 | hCoV-19/England/CAMC-A810C1/2020\|EPI_ISL_610693\|20 | ···················· | ···················· | ···················· | ···················· | ···················· | ···················· |
| O | 4824 | hCoV-19/England/CAMC-A80F6B/2020\|EPI_ISL_610695\|20 | ···················· | ···················· | ···················· | ···················· | ···················· | ···················· |
| O | 4825 | hCoV-19/England/CAMC-A81B39/2020\|EPI_ISL_610696\|20 | ···················· | ···················· | ···················· | ···················· | ···················· | ···················· |
| O | 4826 | hCoV-19/England/CAMC-A821DC/2020\|EPI_ISL_610698\|20 | ···················· | ···················· | ···················· | ···················· | ···················· | ···················· |
| O | 4827 | hCoV-19/England/ALDP-A9277D/2020\|EPI_ISL_611050\|20 | ···················· | ···················· | ···················· | ···················· | ···················· | ···················· |
| O | 4828 | hCoV-19/England/ALDP-A92546/2020\|EPI_ISL_611051\|20 | ···················· | ···················· | ···················· | ···················· | ···················· | ···················· |
| O | 4829 | hCoV-19/England/ALDP-A94061/2020\|EPI_ISL_611052\|20 | ···················· | ···················· | ···················· | ···················· | ···················· | ···················· |
| O | 4830 | hCoV-19/England/ALDP-A92AB0/2020\|EPI_ISL_611053\|20 | ···················· | ···················· | ···················· | ···················· | ···················· | ···················· |
| O | 4831 | hCoV-19/England/ALDP-A928D4/2020\|EPI_ISL_611054\|20 | ···················· | ···················· | ···················· | ···················· | ···················· | ···················· |
| O | 4832 | hCoV-19/England/ALDP-A92E96/2020\|EPI_ISL_611055\|20 | ···················· | ···················· | ···················· | ···················· | ···················· | ···················· |
| O | 4833 | hCoV-19/England/ALDP-A92564/2020\|EPI_ISL_611058\|20 | ···················· | ···················· | ···················· | ···················· | ···················· | ···················· |
| O | 4834 | hCoV-19/England/ALDP-A92E2D/2020\|EPI_ISL_611059\|20 | ···················· | ···················· | ···················· | ···················· | ···················· | ···················· |
| O | 4835 | hCoV-19/England/ALDP-A94878/2020\|EPI_ISL_611060\|20 | ···················· | ···················· | ···················· | ···················· | ···················· | ···················· |
| O | 4836 | hCoV-19/England/ALDP-A94438/2020\|EPI_ISL_611061\|20 | ···················· | ···················· | ···················· | ···················· | ···················· | ···················· |
| O | 4837 | hCoV-19/England/ALDP-A9276E/2020\|EPI_ISL_611062\|20 | ···················· | ···················· | ···················· | ···················· | ···················· | ···················· |
| O | 4838 | hCoV-19/England/ALDP-A93F56/2020\|EPI_ISL_611063\|20 | ···················· | ···················· | ···················· | ···················· | ···················· | ···················· |
| O | 4839 | hCoV-19/England/ALDP-A94887/2020\|EPI_ISL_611064\|20 | ···················· | ···················· | ···················· | ···················· | ···················· | ···················· |
| O | 4840 | hCoV-19/England/ALDP-A9484B/2020\|EPI_ISL_611065\|20 | ···················· | ···················· | ···················· | ···················· | ···················· | ···················· |
| O | 4841 | hCoV-19/England/ALDP-A929E0/2020\|EPI_ISL_611066\|20 | ···················· | ·········A·········· | ···················· | ···················· | ···················· | ···················· |
| O | 4842 | hCoV-19/England/ALDP-A927F5/2020\|EPI_ISL_611068\|20 | ···················· | ···················· | ···················· | ···················· | ···················· | ···················· |
| O | 4843 | hCoV-19/England/ALDP-A93DB6/2020\|EPI_ISL_611069\|20 | ···················· | ···················· | ···················· | ···················· | ···················· | ···················· |
| O | 4844 | hCoV-19/England/CAMB-1B6C7D/2020\|EPI_ISL_611964\|20 | ···················· | ···················· | ···················· | ···················· | ···················· | ···················· |
| O | 4845 | hCoV-19/England/EXET-1380E0/2020\|EPI_ISL_612380\|20 | ···················· | ···················· | ···················· | ···················· | ···················· | ···················· |
| O | 4846 | hCoV-19/England/EXET-1380FF/2020\|EPI_ISL_612381\|20 | ···················· | ···················· | ···················· | ···················· | ···················· | ···················· |
| O | 4847 | hCoV-19/England/EXET-13810B/2020\|EPI_ISL_612382\|20 | ···················· | ···················· | ···················· | ···················· | ···················· | ···················· |
| O | 4848 | hCoV-19/England/EXET-13811A/2020\|EPI_ISL_612383\|20 | ···················· | ···················· | ···················· | ···················· | ···················· | ···················· |
| O | 4849 | hCoV-19/England/EXET-138138/2020\|EPI_ISL_612384\|20 | ···················· | ···················· | ···················· | ···················· | ···················· | ···················· |
| O | 4850 | hCoV-19/England/EXET-138174/2020\|EPI_ISL_612385\|20 | ···················· | ···················· | ···················· | ···················· | ···················· | ···················· |
| O | 4851 | hCoV-19/England/EXET-138183/2020\|EPI_ISL_612386\|20 | ···················· | ···················· | ···················· | ···················· | ···················· | ···················· |
| O | 4852 | hCoV-19/England/EXET-1381B0/2020\|EPI_ISL_612387\|20 | ···················· | ···················· | ···················· | ···················· | ···················· | ···················· |
| O | 4853 | hCoV-19/England/EXET-1381DE/2020\|EPI_ISL_612388\|20 | ···················· | ···················· | ···················· | ···················· | ···················· | ···················· |
| O | 4854 | hCoV-19/England/EXET-1381ED/2020\|EPI_ISL_612389\|20 | ···················· | ···················· | ···················· | ···················· | ···················· | ···················· |
| O | 4856 | hCoV-19/England/EXET-138217/2020\|EPI_ISL_612391\|20 | ···················· | ···················· | ···················· | ···················· | ···················· | ···················· |
| O | 4857 | hCoV-19/England/EXET-138235/2020\|EPI_ISL_612392\|20 | ···················· | ···················· | ···················· | ···················· | ···················· | ···················· |
| O | 4858 | hCoV-19/England/EXET-138244/2020\|EPI_ISL_612393\|20 | ···················· | ···················· | ···················· | ···················· | ···················· | ···················· |
| O | 4859 | hCoV-19/England/EXET-1382CC/2020\|EPI_ISL_612394\|20 | ···················· | ···················· | ···················· | ···················· | ···················· | ···················· |
| O | 4860 | hCoV-19/England/EXET-13836F/2020\|EPI_ISL_612395\|20 | ···················· | ···················· | ···················· | ···················· | ···················· | ···················· |
| O | 4861 | hCoV-19/England/EXET-13838D/2020\|EPI_ISL_612396\|20 | ···················· | ···················· | ···················· | ···················· | ···················· | ···················· |
| O | 4863 | hCoV-19/England/EXET-1383AB/2020\|EPI_ISL_612398\|20 | ···················· | ···················· | ···················· | ···················· | ···················· | ···················· |
| O | 4865 | hCoV-19/England/MILK-A7D1F2/2020\|EPI_ISL_606500\|20 | ···················· | ···················· | ···················· | ···················· | ······T············· | ···················· |
| O | 4866 | hCoV-19/England/MILK-A7C5E8/2020\|EPI_ISL_606501\|20 | ···················· | ···················· | ···················· | ···················· | ····T··············· | ···················· |

| | | | | | | | | |
|---|---|---|---|---|---|---|---|---|
| O | 4867 | hCoV-19/England/MILK-A7C22D/2020\|EPI_ISL_606502\|20 | ···················· | ···················· | ····················· | ···················· | ···················· | ···················· |
| O | 4868 | hCoV-19/England/MILK-A7A6E7/2020\|EPI_ISL_606503\|20 | ···················· | ···················· | ····················· | ···················· | ···················· | ···················· |
| O | 4869 | hCoV-19/England/MILK-A7DF34/2020\|EPI_ISL_606504\|20 | ···················· | ···················· | ····················· | ···················· | ···················· | ···················· |
| O | 4870 | hCoV-19/England/MILK-A7CDA4/2020\|EPI_ISL_606505\|20 | ···················· | ······A············· | ····················· | ···················· | ···················· | ···················· |
| O | 4871 | hCoV-19/England/MILK-A7ADB5/2020\|EPI_ISL_606506\|20 | ···················· | ···················· | ····················· | ···················· | ···················· | ···················· |
| O | 4872 | hCoV-19/England/MILK-A7DD1C/2020\|EPI_ISL_606507\|20 | ···················· | ···················· | ····················· | ···················· | ···················· | ···················· |
| O | 4873 | hCoV-19/England/MILK-A7DD49/2020\|EPI_ISL_606509\|20 | ···················· | ···················· | ····················· | ···················· | ···················· | ···················· |
| O | 4874 | hCoV-19/England/MILK-A7C74C/2020\|EPI_ISL_606510\|20 | ···················· | ···················· | ····················· | ···················· | ···················· | ···················· |
| O | 4875 | hCoV-19/England/MILK-A7AF37/2020\|EPI_ISL_606511\|20 | ···················· | ···················· | ····················· | ···················· | ···················· | ···················· |
| O | 4876 | hCoV-19/England/MILK-A7D3CF/2020\|EPI_ISL_606513\|20 | ···················· | ···················· | ····················· | ···················· | ···················· | ···················· |
| O | 4877 | hCoV-19/England/MILK-A7AA72/2020\|EPI_ISL_606514\|20 | ···················· | ·A·················· | ····················· | ···················· | ···················· | ···A················ |
| O | 4878 | hCoV-19/England/MILK-A7CA43/2020\|EPI_ISL_606515\|20 | ···················· | ···················· | ····················· | ···················· | ···················· | ···················· |
| O | 4879 | hCoV-19/England/MILK-A7D13E/2020\|EPI_ISL_606517\|20 | ···················· | ·A·················· | ····················· | ···················· | ···················· | ···A················ |
| O | 4880 | hCoV-19/England/MILK-A7D514/2020\|EPI_ISL_606518\|20 | ···················· | ···················· | ····················· | ···················· | ···················· | ···················· |
| O | 4881 | hCoV-19/England/MILK-A7D875/2020\|EPI_ISL_606519\|20 | ···················· | ···················· | ····················· | ···················· | ···················· | ···················· |
| O | 4882 | hCoV-19/England/MILK-A7D9CD/2020\|EPI_ISL_606520\|20 | ···················· | ···················· | ····················· | ···················· | ···················· | ···················· |
| O | 4883 | hCoV-19/England/MILK-A7C463/2020\|EPI_ISL_606522\|20 | ···················· | ···················· | ····················· | ···················· | ···················· | ···················· |
| O | 4884 | hCoV-19/England/MILK-A7AE2B/2020\|EPI_ISL_606525\|20 | ···················· | ···················· | ····················· | ···················· | ···················· | ···················· |
| O | 4885 | hCoV-19/England/MILK-A7D81B/2020\|EPI_ISL_606526\|20 | ···················· | C··················· | ····················· | ···················· | ···················· | ···················· |
| O | 4886 | hCoV-19/England/MILK-A7A83C/2020\|EPI_ISL_606527\|20 | ···················· | ···················· | ····················· | ···················· | ···················· | ···················· |
| O | 4887 | hCoV-19/England/MILK-A7C64F/2020\|EPI_ISL_606528\|20 | ···················· | ···················· | ····················· | ···················· | ···················· | ···················· |
| O | 4888 | hCoV-19/England/MILK-A7AABE/2020\|EPI_ISL_606529\|20 | ···················· | ···················· | ····················· | ···················· | ···················· | ···················· |
| O | 4889 | hCoV-19/England/MILK-A7A887/2020\|EPI_ISL_606540\|20 | ···················· | ···················· | ····················· | ···················· | ···················· | ···················· |
| O | 4890 | hCoV-19/England/MILK-A7A77B/2020\|EPI_ISL_606541\|20 | ···················· | ···················· | ····················· | ···················· | ···················· | ···················· |
| O | 4891 | hCoV-19/England/MILK-A7DE0A/2020\|EPI_ISL_606542\|20 | ···················· | ···················· | ····················· | ···················· | ···················· | ···················· |
| O | 4892 | hCoV-19/England/MILK-A7A84B/2020\|EPI_ISL_606544\|20 | ···················· | ···················· | ····················· | ···················· | ···················· | ···················· |
| O | 4893 | hCoV-19/England/MILK-A7DF07/2020\|EPI_ISL_606545\|20 | ···················· | ···················· | ····················· | ···················· | ···················· | ···················· |
| O | 4894 | hCoV-19/England/MILK-A7D884/2020\|EPI_ISL_606546\|20 | ···················· | ···················· | ····················· | ···················· | ···················· | ···················· |
| O | 4895 | hCoV-19/England/MILK-A7D365/2020\|EPI_ISL_606548\|20 | ···················· | ·A·················· | ····················· | ···················· | ···················· | ···A················ |
| O | 4896 | hCoV-19/England/MILK-A7D7E1/2020\|EPI_ISL_606550\|20 | ···················· | ·A·················· | ····················· | ···················· | ···················· | ···················· |
| O | 4897 | hCoV-19/England/MILK-A7D268/2020\|EPI_ISL_606551\|20 | ···················· | ···················· | ····················· | ···················· | ···················· | ···················· |
| O | 4898 | hCoV-19/England/MILK-A7C31B/2020\|EPI_ISL_606552\|20 | ···················· | ···················· | ····················· | ···················· | ···················· | ···················· |
| O | 4899 | hCoV-19/England/MILK-A7CB6E/2020\|EPI_ISL_606553\|20 | ···················· | ···················· | ····················· | ···················· | ···················· | ···················· |
| O | 4900 | hCoV-19/England/MILK-A7CDC2/2020\|EPI_ISL_606554\|20 | ···················· | ···················· | ····················· | ···················· | ······T············· | ···················· |
| O | 4901 | hCoV-19/England/MILK-A7DCA6/2020\|EPI_ISL_606555\|20 | ···················· | ···················· | ····················· | ···················· | ···················· | ···················· |
| O | 4902 | hCoV-19/England/MILK-A7D2FF/2020\|EPI_ISL_606556\|20 | ···················· | ···················· | ····················· | ···················· | ···················· | ···················· |
| O | 4903 | hCoV-19/England/MILK-A7C24B/2020\|EPI_ISL_606557\|20 | ···················· | ···················· | ····················· | ···················· | ···················· | ···················· |
| O | 4904 | hCoV-19/England/MILK-A7D286/2020\|EPI_ISL_606558\|20 | ···················· | ···················· | ····················· | ···················· | ···················· | ···················· |
| O | 4905 | hCoV-19/England/MILK-A7DB30/2020\|EPI_ISL_606559\|20 | ···················· | ···················· | ····················· | ···················· | ···················· | ···················· |
| O | 4906 | hCoV-19/England/MILK-A7DA8E/2020\|EPI_ISL_606560\|20 | ···················· | ···················· | ····················· | ···················· | ······T············· | ···················· |
| O | 4907 | hCoV-19/England/MILK-A7D5E7/2020\|EPI_ISL_606561\|20 | ···················· | ···················· | ····················· | ···················· | ···················· | ···················· |
| O | 4908 | hCoV-19/England/MILK-A7C630/2020\|EPI_ISL_606562\|20 | ···················· | ···················· | ····················· | ···················· | ···················· | ···················· |
| O | 4909 | hCoV-19/England/MILK-A7D1D4/2020\|EPI_ISL_606563\|20 | ···················· | ···················· | ····················· | ···················· | ···················· | ···················· |
| O | 4910 | hCoV-19/England/MILK-A7CDB3/2020\|EPI_ISL_606564\|20 | ···················· | ···················· | ····················· | ···················· | ···················· | ···················· |
| O | 4911 | hCoV-19/England/MILK-A7DB12/2020\|EPI_ISL_606566\|20 | ···················· | ···················· | ····················· | ···················· | ···················· | ···················· |
| O | 4912 | hCoV-19/England/MILK-A7AAFA/2020\|EPI_ISL_606567\|20 | ···················· | ···················· | ····················· | ···················· | ···················· | ···················· |
| O | 4913 | hCoV-19/England/MILK-A7C71F/2020\|EPI_ISL_606568\|20 | ···················· | ···················· | ····················· | ···················· | ···················· | ···················· |
| O | 4914 | hCoV-19/England/MILK-A7D2C2/2020\|EPI_ISL_606569\|20 | ···················· | ···················· | ····················· | ···················· | ···················· | ···················· |
| O | 4915 | hCoV-19/England/MILK-A7DB21/2020\|EPI_ISL_606570\|20 | ···················· | ···················· | ····················· | ···················· | ···················· | ···················· |
| O | 4916 | hCoV-19/England/MILK-A7C66D/2020\|EPI_ISL_606571\|20 | ···················· | ···················· | ····················· | ···················· | ······T············· | ···················· |
| O | 4917 | hCoV-19/England/MILK-A7C436/2020\|EPI_ISL_606572\|20 | ···················· | ···················· | ····················· | ···················· | ···················· | ···················· |
| O | 4918 | hCoV-19/England/MILK-A7C269/2020\|EPI_ISL_606574\|20 | ···················· | ···················· | ···T················· | ···················· | ···················· | ···················· |
| O | 4919 | hCoV-19/England/MILK-A7D4EA/2020\|EPI_ISL_606575\|20 | ···················· | ·A·················· | ····················· | ···················· | ···················· | ···A················ |
| O | 4920 | hCoV-19/England/MILK-A7D787/2020\|EPI_ISL_606576\|20 | ···················· | ···················· | ····················· | ···················· | ···················· | ···················· |
| O | 4921 | hCoV-19/England/MILK-A7D435/2020\|EPI_ISL_606577\|20 | ···················· | ···················· | ····················· | ···················· | ···················· | ···················· |
| O | 4922 | hCoV-19/England/MILK-A7D5AB/2020\|EPI_ISL_606578\|20 | ···················· | ·A·················· | ····················· | ···················· | ···················· | ···················· |
| O | 4923 | hCoV-19/England/MILK-A7D66C/2020\|EPI_ISL_606579\|20 | ···················· | ···················· | ····················· | ···················· | ···················· | ···················· |
| O | 4924 | hCoV-19/England/ALDP-A6DF91/2020\|EPI_ISL_607800\|20 | ···················· | ···················· | ····················· | ···················· | ···················· | ···················· |
| O | 4925 | hCoV-19/England/ALDP-A6E4CE/2020\|EPI_ISL_607807\|20 | ···················· | ···················· | ····················· | ···················· | ···················· | ···················· |
| O | 4926 | hCoV-19/England/ALDP-A70757/2020\|EPI_ISL_607808\|20 | ···················· | ···················· | ····················· | ···················· | ···················· | ···················· |
| O | 4927 | hCoV-19/England/ALDP-A6DA45/2020\|EPI_ISL_607809\|20 | ···················· | ···················· | ····················· | ···················· | ···················· | ···················· |
| O | 4928 | hCoV-19/England/ALDP-A6D869/2020\|EPI_ISL_607810\|20 | ···················· | ···················· | ····················· | ···················· | ···················· | ···················· |
| O | 4929 | hCoV-19/England/ALDP-A6E1E5/2020\|EPI_ISL_607811\|20 | ···················· | ···················· | ····················· | ···················· | ···················· | ···················· |
| O | 4930 | hCoV-19/England/ALDP-A6DA90/2020\|EPI_ISL_607812\|20 | ···················· | ···················· | ····················· | ···················· | ···················· | ···················· |
| O | 4931 | hCoV-19/England/ALDP-A7058A/2020\|EPI_ISL_607813\|20 | ···················· | ···················· | ····················· | ···················· | ···················· | ···················· |
| O | 4932 | hCoV-19/England/ALDP-A6F68B/2020\|EPI_ISL_607814\|20 | ···················· | ···················· | ····················· | ···················· | ···················· | ···················· |
| O | 4933 | hCoV-19/England/ALDP-A6E507/2020\|EPI_ISL_607816\|20 | ···················· | ···················· | ····················· | ···················· | ···················· | ···················· |
| O | 4934 | hCoV-19/England/ALDP-A6E7E3/2020\|EPI_ISL_607817\|20 | ···················· | ···················· | ····················· | ···················· | ···················· | ···················· |
| O | 4935 | hCoV-19/England/ALDP-A6ED78/2020\|EPI_ISL_607818\|20 | ···················· | ···················· | ····················· | ···················· | ···················· | ···················· |
| O | 4936 | hCoV-19/England/ALDP-A6E7A7/2020\|EPI_ISL_607819\|20 | ···················· | ···················· | ····················· | ···················· | ···················· | ···················· |
| O | 4937 | hCoV-19/England/ALDP-A6E051/2020\|EPI_ISL_607820\|20 | ···················· | ···················· | ····················· | ···················· | ···················· | ···················· |
| O | 4938 | hCoV-19/England/ALDP-A6DE94/2020\|EPI_ISL_607821\|20 | ···················· | ···················· | ····················· | ···················· | ···················· | ···················· |
| O | 4939 | hCoV-19/England/ALDP-A70599/2020\|EPI_ISL_607822\|20 | ···················· | ···················· | ····················· | ···················· | ···················· | ···················· |
| O | 4940 | hCoV-19/England/ALDP-A6F6E5/2020\|EPI_ISL_607823\|20 | ···················· | ·········T·········· | ····················· | ···················· | ···················· | ···················· |
| O | 4941 | hCoV-19/England/ALDP-A6EA80/2020\|EPI_ISL_607825\|20 | ···················· | ···················· | ····················· | ···················· | ···················· | ···················· |
| O | 4942 | hCoV-19/England/ALDP-A6ED2D/2020\|EPI_ISL_607826\|20 | ···················· | ···················· | ····················· | ···················· | ···················· | ···················· |
| O | 4943 | hCoV-19/England/ALDP-A70423/2020\|EPI_ISL_607827\|20 | ···················· | ···················· | ····················· | ···················· | ···················· | ···················· |
| O | 4944 | hCoV-19/England/ALDP-A6E561/2020\|EPI_ISL_607830\|20 | ···················· | ···················· | ····················· | ···················· | ···················· | ···················· |
| O | 4945 | hCoV-19/England/ALDP-A6E76B/2020\|EPI_ISL_607831\|20 | ···················· | ···················· | ····················· | ···················· | ···················· | ···················· |
| O | 4946 | hCoV-19/England/ALDP-A6FA34/2020\|EPI_ISL_607832\|20 | ···················· | ···················· | ····················· | ···················· | ···················· | ···················· |

O 4947 hCoV-19/England/ALDP-A6D84B/2020|EPI_ISL_607833|20 ···················· ···················· ····················· ···················· ···················· ····················
O 4948 hCoV-19/England/ALDP-A6E80E/2020|EPI_ISL_607835|20 ···················· ···················· ····················· ···················· ···················· ····················
O 4949 hCoV-19/England/ALDP-A6ED4B/2020|EPI_ISL_607836|20 ···················· ···················· ····················· ···················· ···················· ····················
O 4950 hCoV-19/England/ALDP-A6DC21/2020|EPI_ISL_607838|20 ···················· ···················· ····················· ···················· ···················· ····················
O 4951 hCoV-19/England/ALDP-A6FA61/2020|EPI_ISL_607839|20 ···················· ···················· ····················· ···················· ···················· ····················
O 4952 hCoV-19/England/ALDP-A6E8D1/2020|EPI_ISL_607840|20 ···················· ···················· ····················· ···················· ···················· ····················
O 4953 hCoV-19/England/ALDP-A6DFA0/2020|EPI_ISL_607841|20 ···················· ···················· ····················· ···················· ·····T·············· ····················
O 4954 hCoV-19/England/ALDP-A6F9CE/2020|EPI_ISL_607844|20 ···················· ···················· ····················· ···················· ···················· ····················
O 4955 hCoV-19/England/ALDP-A704C9/2020|EPI_ISL_607846|20 ···················· ···················· ····················· ···················· ···················· ····················
O 4956 hCoV-19/England/ALDP-A6D91B/2020|EPI_ISL_607847|20 ···················· ···················· ····················· ···················· ···················· ····················
O 4957 hCoV-19/England/ALDP-A706D2/2020|EPI_ISL_607848|20 ···················· ···················· ····················· ···················· ···················· ····················
O 4958 hCoV-19/England/ALDP-A6E82C/2020|EPI_ISL_607849|20 ···················· ···················· ····················· ···················· ···················· ····················
O 4959 hCoV-19/England/ALDP-A6DF19/2020|EPI_ISL_607850|20 ···················· ···················· ····················· ···················· ···················· ····················
O 4960 hCoV-19/England/ALDP-A6DB24/2020|EPI_ISL_607852|20 ···················· ···················· ····················· ···················· ···················· ····················
O 4961 hCoV-19/England/ALDP-A70511/2020|EPI_ISL_607853|20 ···················· ···················· ····················· ···················· ···················· ····················
O 4962 hCoV-19/England/ALDP-A6E947/2020|EPI_ISL_607854|20 ···················· ···················· ····················· ···················· ···················· ····················
O 4963 hCoV-19/England/ALDP-A706E1/2020|EPI_ISL_607855|20 ···················· ···················· ····················· ···················· ···················· ··A·················
O 4964 hCoV-19/England/ALDP-A705B7/2020|EPI_ISL_607856|20 ···················· ···················· ····················· ···················· ···················· ····················
O 4965 hCoV-19/England/ALDP-A6E77A/2020|EPI_ISL_607859|20 ···················· ···················· ····················· ···················· ···················· ····················
O 4966 hCoV-19/England/ALDP-A6F0AB/2020|EPI_ISL_607870|20 ···················· ···················· ····················· ···················· ···················· ····················
O 4967 hCoV-19/England/ALDP-A6DB51/2020|EPI_ISL_607871|20 ···················· ···················· ····················· ···················· ···················· ····················
O 4968 hCoV-19/England/ALDP-A6DA81/2020|EPI_ISL_607872|20 ···················· ···················· ····················· ···················· ···················· ····················
O 4969 hCoV-19/England/ALDP-A6E1F4/2020|EPI_ISL_607873|20 ···················· ···················· ····················· ···················· ···················· ····················
O 4970 hCoV-19/England/ALDP-A6E419/2020|EPI_ISL_607874|20 ···················· ···················· ····················· ···················· ···················· ····················
O 4971 hCoV-19/England/ALDP-A6DBCA/2020|EPI_ISL_607875|20 ···················· ···················· ····················· ···················· ···················· ····················
O 4972 hCoV-19/England/ALDP-A6EF81/2020|EPI_ISL_607876|20 ···················· ···················· ····················· ···················· ···················· ····················
O 4973 hCoV-19/England/ALDP-A6ED69/2020|EPI_ISL_607877|20 ···················· ···················· ····················· ···················· ···················· ····················
O 4974 hCoV-19/England/ALDP-A6DAFA/2020|EPI_ISL_607878|20 ···················· ···················· ····················· ···················· ···················· ····················
O 4975 hCoV-19/England/ALDP-A6EF36/2020|EPI_ISL_607879|20 ···················· ···················· ····················· ···················· ···················· ····················
O 4976 hCoV-19/England/ALDP-A6DA72/2020|EPI_ISL_607880|20 ···················· ···················· ····················· ···················· ···················· ····················
O 4977 hCoV-19/England/ALDP-A6F08D/2020|EPI_ISL_607881|20 ···················· ···················· ····················· ···················· ···················· ····················
O 4978 hCoV-19/England/ALDP-A6E006/2020|EPI_ISL_607882|20 ···················· ···················· ····················· ···················· ···················· ····················
O 4979 hCoV-19/England/ALDP-A6F5BB/2020|EPI_ISL_607883|20 ···················· ···················· ····················· ···················· ···················· ····················
O 4980 hCoV-19/England/ALDP-A6D92A/2020|EPI_ISL_607884|20 ···················· ···················· ····················· ···················· ···················· ····················
O 4981 hCoV-19/England/ALDP-A7071B/2020|EPI_ISL_607885|20 ···················· ···················· ····················· ···················· ···················· ····················
O 4982 hCoV-19/England/ALDP-A6DBF7/2020|EPI_ISL_607886|20 ···················· ···················· ····················· ···················· ···················· ····················
O 4983 hCoV-19/England/ALDP-A6DF37/2020|EPI_ISL_607887|20 ···················· ···················· ····················· ···················· ···················· ····················
O 4984 hCoV-19/England/ALDP-A6F427/2020|EPI_ISL_607888|20 ···················· ···················· ····················· ···················· ···················· ····················
O 4985 hCoV-19/England/ALDP-A6DEEF/2020|EPI_ISL_607889|20 ···················· ···················· ····················· ···················· ···················· ····················
O 4986 hCoV-19/England/ALDP-A6F9DD/2020|EPI_ISL_607890|20 ···················· ···················· ····················· ···················· ···················· ····················
O 4987 hCoV-19/England/ALDP-A6F0C9/2020|EPI_ISL_607891|20 ···················· ···················· ····················· ···················· ···················· ····················
O 4988 hCoV-19/England/ALDP-A6DFBF/2020|EPI_ISL_607892|20 ···················· ···················· ····················· ···················· ···················· ····················
O 4989 hCoV-19/England/ALDP-A6F603/2020|EPI_ISL_607895|20 ···················· ···················· ····················· ···················· ···················· ····················
O 4990 hCoV-19/England/ALDP-A6F524/2020|EPI_ISL_607897|20 ···················· ···················· ····················· ···················· ···················· ····················
O 4991 hCoV-19/England/ALDP-A92959/2020|EPI_ISL_611008|20 ···················· ···················· ····················· ···················· ···················· ····················
O 4992 hCoV-19/England/ALDP-A935AF/2020|EPI_ISL_611009|20 ···················· ···················· ····················· ···················· ···················· ····················
O 4993 hCoV-19/England/ALDP-A94386/2020|EPI_ISL_611010|20 ···················· ···················· ····················· ···················· ···················· ····················
O 4994 hCoV-19/England/ALDP-A93D5C/2020|EPI_ISL_611011|20 ···················· ···················· ····················· ···················· ···················· ····················
O 4995 hCoV-19/England/ALDP-A96E74/2020|EPI_ISL_611012|20 ···················· ···················· ····················· ···················· ···················· ····················
O 4996 hCoV-19/England/ALDP-A943D1/2020|EPI_ISL_611013|20 ···················· ···················· ····················· ···················· ···················· ····················
O 4997 hCoV-19/England/ALDP-A935EB/2020|EPI_ISL_611016|20 ···················· ···················· ····················· ···················· ···················· ····················
O 4998 hCoV-19/England/ALDP-A93BF8/2020|EPI_ISL_611017|20 ···················· ···················· ····················· ···················· ···················· ····················
O 4999 hCoV-19/England/ALDP-A929FF/2020|EPI_ISL_611018|20 ···················· ···················· ····················· ···················· ···················· ····················
O 5000 hCoV-19/England/ALDP-A92A0B/2020|EPI_ISL_611019|20 ···················· ···················· ····················· ···················· ···················· ····················
O 5001 hCoV-19/England/ALDP-A92670/2020|EPI_ISL_611020|20 ···················· ···················· ····················· ···················· ···················· ····················
O 5002 hCoV-19/England/ALDP-A92582/2020|EPI_ISL_611021|20 ···················· ·A·················· ····················· ···················· ···················· ····················
O 5003 hCoV-19/England/ALDP-A93CF5/2020|EPI_ISL_611022|20 ···················· ·A·················· ····················· ···················· ···················· ··A·················
O 5004 hCoV-19/England/ALDP-A93660/2020|EPI_ISL_611023|20 ···················· ···················· ····················· ···················· ···················· ····················
O 5005 hCoV-19/England/ALDP-A9482D/2020|EPI_ISL_611024|20 ···················· ···················· ····················· ···················· ···················· ····················
O 5006 hCoV-19/England/ALDP-A92388/2020|EPI_ISL_611025|20 ···················· ···················· ····················· ···················· ···················· ····················
O 5007 hCoV-19/England/ALDP-A92A1A/2020|EPI_ISL_611026|20 ···················· ···················· ····················· ···················· ···················· ····················
O 5008 hCoV-19/England/ALDP-A940AD/2020|EPI_ISL_611027|20 ···················· ···················· ····················· ···················· ···················· ····················
O 5009 hCoV-19/England/ALDP-A92CF6/2020|EPI_ISL_611028|20 ···················· ···················· ····················· ···················· ···················· ····················
O 5010 hCoV-19/England/ALDP-A9385B/2020|EPI_ISL_611029|20 ···················· ···················· ····················· ···················· ···················· ····················
O 5011 hCoV-19/England/ALDP-A9467E/2020|EPI_ISL_611030|20 ···················· ···················· ····················· ···················· ···················· ····················
O 5012 hCoV-19/England/ALDP-A92528/2020|EPI_ISL_611031|20 ···················· ·A·················· ····················· ···················· ···················· ··A·················
O 5013 hCoV-19/England/ALDP-A927D7/2020|EPI_ISL_611032|20 ···················· ···················· ····················· ···················· ···················· ····················
O 5014 hCoV-19/England/ALDP-A9243A/2020|EPI_ISL_611033|20 ···················· ···················· ····················· ···················· ···················· ····················
O 5015 hCoV-19/England/ALDP-A92E87/2020|EPI_ISL_611034|20 ···················· ···················· ····················· ···················· ···················· ····················
O 5016 hCoV-19/England/ALDP-A92C7E/2020|EPI_ISL_611035|20 ···················· ···················· ····················· ···················· ···················· ····················
O 5017 hCoV-19/England/ALDP-A92704/2020|EPI_ISL_611036|20 ···················· ···················· ····················· ···················· ···················· ····················
O 5018 hCoV-19/England/ALDP-A9480F/2020|EPI_ISL_611038|20 ···················· ···················· ····················· ···················· ···················· ····················
O 5019 hCoV-19/England/ALDP-A93ADD/2020|EPI_ISL_611039|20 ···················· ···················· ····················· ···················· ···················· ····················
O 5020 hCoV-19/England/ALDP-A9386A/2020|EPI_ISL_611041|20 ···················· ···················· ····················· ···················· ···················· ····················
O 5021 hCoV-19/England/ALDP-A9242B/2020|EPI_ISL_611042|20 ···················· ···················· ····················· ···················· ···················· ····················
O 5022 hCoV-19/England/ALDP-A92C50/2020|EPI_ISL_611043|20 ···················· ···················· ····················· ···················· ···················· ····················
O 5023 hCoV-19/England/ALDP-A93581/2020|EPI_ISL_611044|20 ···················· ···················· ····················· ···················· ···················· ····················
O 5024 hCoV-19/England/ALDP-A93958/2020|EPI_ISL_611046|20 ···················· ···················· ····················· ···················· ···················· ····················
O 5025 hCoV-19/England/ALDP-A93BBC/2020|EPI_ISL_611047|20 ···················· ···················· ····················· ···················· ···················· ····················
O 5026 hCoV-19/England/ALDP-A94007/2020|EPI_ISL_611049|20 ···················· ···················· ····················· ···················· ···················· ····················

| | | | | | | | | |
|---|---|---|---|---|---|---|---|---|
| O | 5027 | hCoV-19/England/ALDP-A92A92/2020\|EPI_ISL_611070\|20 | ···················· | ···················· | ····················· | ···················· | ···················· | ···················· |
| O | 5028 | hCoV-19/England/ALDP-A96E1A/2020\|EPI_ISL_611071\|20 | ···················· | ···················· | ····················· | ···················· | ···················· | ···················· |
| O | 5029 | hCoV-19/England/ALDP-A94896/2020\|EPI_ISL_611073\|20 | ···················· | ···················· | ····················· | ···················· | ···················· | ···················· |
| O | 5030 | hCoV-19/England/ALDP-A92652/2020\|EPI_ISL_611074\|20 | ···················· | ···················· | ····················· | ···················· | ···················· | ···················· |
| O | 5031 | hCoV-19/England/ALDP-A93DD4/2020\|EPI_ISL_611075\|20 | ···················· | ···················· | ····················· | ···················· | ···················· | ···················· |
| O | 5032 | hCoV-19/England/ALDP-A93D10/2020\|EPI_ISL_611076\|20 | ···················· | ···················· | ····················· | ···················· | ···················· | ···················· |
| O | 5033 | hCoV-19/England/ALDP-A9235B/2020\|EPI_ISL_611077\|20 | ···················· | ···················· | ····················· | ···················· | ···················· | ···················· |
| O | 5034 | hCoV-19/England/ALDP-A92519/2020\|EPI_ISL_611078\|20 | ···················· | ···················· | ····················· | ···················· | ···················· | ···················· |
| O | 5035 | hCoV-19/England/ALDP-A93A82/2020\|EPI_ISL_611079\|20 | ···················· | ···················· | ····················· | ···················· | ···················· | ···················· |
| O | 5036 | hCoV-19/England/ALDP-A93554/2020\|EPI_ISL_611080\|20 | ···················· | ···················· | ····················· | ···················· | ···················· | ···················· |
| O | 5037 | hCoV-19/England/ALDP-A93E77/2020\|EPI_ISL_611081\|20 | ···················· | ···················· | ····················· | ···················· | ···················· | ···················· |
| O | 5038 | hCoV-19/England/ALDP-A92BCC/2020\|EPI_ISL_611082\|20 | ···················· | ···················· | ····················· | ···················· | ···················· | ···················· |
| O | 5039 | hCoV-19/England/ALDP-A93AEC/2020\|EPI_ISL_611084\|20 | ···················· | ···················· | ····················· | ···················· | ···················· | ···················· |
| O | 5040 | hCoV-19/England/ALDP-A96EED/2020\|EPI_ISL_611085\|20 | ···················· | ···················· | ····················· | ···················· | ···················· | ···················· |
| O | 5041 | hCoV-19/England/ALDP-A923C4/2020\|EPI_ISL_611086\|20 | ···················· | ···················· | ····················· | ···················· | ···················· | ···················· |
| O | 5042 | hCoV-19/England/ALDP-A92B08/2020\|EPI_ISL_611087\|20 | ···················· | ···················· | ····················· | ···················· | ···················· | ···················· |
| O | 5043 | hCoV-19/England/ALDP-A93EFF/2020\|EPI_ISL_611089\|20 | ···················· | ·A·················· | ····················· | ···················· | ···················· | ··A················· |
| O | 5044 | hCoV-19/England/ALDP-A93624/2020\|EPI_ISL_611090\|20 | ···················· | ···················· | ····················· | ···················· | ···················· | ···················· |
| O | 5045 | hCoV-19/England/ALDP-A92537/2020\|EPI_ISL_611091\|20 | ···················· | ···················· | ····················· | ···················· | ···················· | ···················· |
| O | 5046 | hCoV-19/England/ALDP-A94025/2020\|EPI_ISL_611092\|20 | ···················· | ···················· | ····················· | ···················· | ···················· | ···················· |
| O | 5047 | hCoV-19/England/ALDP-A93E3B/2020\|EPI_ISL_611095\|20 | ···················· | ···················· | ····················· | ···················· | ···················· | ···················· |
| O | 5048 | hCoV-19/England/ALDP-A92CAB/2020\|EPI_ISL_611096\|20 | ···················· | ···················· | ····················· | ···················· | ···················· | ···················· |
| O | 5049 | hCoV-19/England/ALDP-A93FA1/2020\|EPI_ISL_611097\|20 | ···················· | ···················· | ····················· | ···················· | ···················· | ···················· |
| O | 5050 | hCoV-19/Scotland/EDB8351/2020\|EPI_ISL_612339\|2020- | ···················· | ···················· | ····················· | ···················· | ···················· | ···················· |
| O | 5051 | hCoV-19/Scotland/EDB8352/2020\|EPI_ISL_612340\|2020- | ···················· | ···················· | ····················· | ···················· | ···················· | ···················· |
| O | 5052 | hCoV-19/Scotland/EDB8360/2020\|EPI_ISL_612341\|2020- | ···················· | ···················· | ····················· | ···················· | ···················· | ···················· |
| O | 5053 | hCoV-19/Scotland/EDB8361/2020\|EPI_ISL_612342\|2020- | ···················· | ···················· | ····················· | ···················· | ···················· | ···················· |
| O | 5054 | hCoV-19/Scotland/EDB8366/2020\|EPI_ISL_612343\|2020- | ···················· | ···················· | ····················· | ···················· | ···················· | ···················· |
| O | 5055 | hCoV-19/Scotland/EDB8368/2020\|EPI_ISL_612344\|2020- | ···················· | ···················· | ····················· | ···················· | ···················· | ···················· |
| O | 5056 | hCoV-19/Scotland/EDB8369/2020\|EPI_ISL_612345\|2020- | ···················· | ···················· | ····················· | ···················· | ···················· | ···················· |
| O | 5057 | hCoV-19/Scotland/EDB8370/2020\|EPI_ISL_612346\|2020- | ···················· | ···················· | ····················· | ···················· | ···················· | ···················· |
| O | 5058 | hCoV-19/Scotland/EDB8371/2020\|EPI_ISL_612347\|2020- | ···················· | ···················· | ····················· | ···················· | ···················· | ···················· |
| O | 5059 | hCoV-19/Scotland/EDB8377/2020\|EPI_ISL_612348\|2020- | ···················· | ···················· | ····················· | ···················· | ···················· | ···················· |
| O | 5060 | hCoV-19/Scotland/EDB8378/2020\|EPI_ISL_612349\|2020- | ···················· | ···················· | ····················· | ···················· | ···················· | ···················· |
| O | 5061 | hCoV-19/Scotland/EDB8381/2020\|EPI_ISL_612350\|2020- | ···················· | ···················· | ····················· | ···················· | ···················· | ···················· |
| O | 5062 | hCoV-19/Scotland/EDB8382/2020\|EPI_ISL_612351\|2020- | ···················· | ···················· | ····················· | ···················· | ···················· | ···················· |
| O | 5064 | hCoV-19/Scotland/EDB8404/2020\|EPI_ISL_612353\|2020- | ···················· | ···················· | ····················· | ···················· | ···················· | ···················· |
| O | 5065 | hCoV-19/Scotland/EDB8405/2020\|EPI_ISL_612354\|2020- | ···················· | ···················· | ····················· | ···················· | ···················· | ···················· |
| O | 5067 | hCoV-19/Scotland/EDB8415/2020\|EPI_ISL_612356\|2020- | ···················· | ···················· | ····················· | ···················· | ···················· | ···················· |
| O | 5068 | hCoV-19/Scotland/EDB8431/2020\|EPI_ISL_612357\|2020- | ···················· | ···················· | ····················· | ···················· | ···················· | ···················· |
| O | 5069 | hCoV-19/Scotland/EDB8439/2020\|EPI_ISL_612358\|2020- | ···················· | ···················· | ····················· | ···················· | ···················· | ···················· |
| O | 5070 | hCoV-19/Scotland/EDB8440/2020\|EPI_ISL_612359\|2020- | ···················· | ···················· | ····················· | ···················· | ···················· | ···················· |
| O | 5071 | hCoV-19/Scotland/EDB8441/2020\|EPI_ISL_612360\|2020- | ···················· | ···················· | ····················· | ···················· | ···················· | ···················· |
| O | 5072 | hCoV-19/Scotland/EDB8442/2020\|EPI_ISL_612361\|2020- | ···················· | ···················· | ····················· | ···················· | ···················· | ···················· |
| O | 5073 | hCoV-19/Scotland/EDB8443/2020\|EPI_ISL_612362\|2020- | ···················· | ···················· | ····················· | ···················· | ···················· | ···················· |
| O | 5074 | hCoV-19/Scotland/EDB8451/2020\|EPI_ISL_612363\|2020- | ···················· | ···················· | ····················· | ···················· | ···················· | ···················· |
| O | 5075 | hCoV-19/Scotland/EDB8452/2020\|EPI_ISL_612364\|2020- | ···················· | ···················· | ····················· | ···················· | ···················· | ···················· |
| O | 5076 | hCoV-19/Scotland/EDB8455/2020\|EPI_ISL_612365\|2020- | ···················· | ···················· | ····················· | ···················· | ···················· | ···················· |
| O | 5077 | hCoV-19/Scotland/EDB8457/2020\|EPI_ISL_612366\|2020- | ···················· | ···················· | ····················· | ···················· | ···················· | ···················· |
| O | 5078 | hCoV-19/Scotland/EDB8459/2020\|EPI_ISL_612367\|2020- | ···················· | ···················· | ····················· | ···················· | ···················· | ···················· |
| O | 5080 | hCoV-19/Scotland/EDB8476/2020\|EPI_ISL_612369\|2020- | ···················· | ···················· | ····················· | ···················· | ···················· | ···················· |
| O | 5081 | hCoV-19/Scotland/EDB8486/2020\|EPI_ISL_612370\|2020- | ···················· | ···················· | ····················· | ···················· | ···················· | ···················· |
| O | 5082 | hCoV-19/Scotland/EDB8487/2020\|EPI_ISL_612371\|2020- | ···················· | ···················· | ····················· | ···················· | ···················· | ···················· |
| O | 5083 | hCoV-19/Scotland/EDB8490/2020\|EPI_ISL_612372\|2020- | ···················· | ·········A·········· | ····················· | ···················· | ···················· | ···················· |
| O | 5084 | hCoV-19/Scotland/EDB8491/2020\|EPI_ISL_612373\|2020- | ···················· | ···················· | ····················· | ···················· | ···················· | ···················· |
| O | 5085 | hCoV-19/Scotland/EDB8493/2020\|EPI_ISL_612374\|2020- | ···················· | ···················· | ····················· | ···················· | ···················· | ···················· |
| O | 5086 | hCoV-19/England/EXET-13802C/2020\|EPI_ISL_612375\|20 | ···················· | ···················· | ····················· | ···················· | ···················· | ···················· |
| O | 5087 | hCoV-19/England/EXET-13803B/2020\|EPI_ISL_612376\|20 | ···················· | ···················· | ····················· | ···················· | ···················· | ···················· |
| O | 5088 | hCoV-19/England/EXET-13804A/2020\|EPI_ISL_612377\|20 | ···················· | ···················· | ····················· | ···················· | ···················· | ···················· |
| O | 5089 | hCoV-19/England/EXET-138059/2020\|EPI_ISL_612378\|20 | ···················· | ···················· | ····················· | ···················· | ···················· | ···················· |
| O | 5090 | hCoV-19/England/EXET-138068/2020\|EPI_ISL_612379\|20 | ···················· | ···················· | ····················· | ···················· | ···················· | ···················· |
| O | 5091 | hCoV-19/Gibraltar/204421209/2020\|EPI_ISL_613670\|20 | ···················· | ···················· | ····················· | ···················· | ···················· | ···················· |
| O | 5092 | hCoV-19/Gibraltar/204421210/2020\|EPI_ISL_613671\|20 | ···················· | ···················· | ····················· | ···················· | ···················· | ···················· |
| O | 5093 | hCoV-19/Gibraltar/204421211/2020\|EPI_ISL_613672\|20 | ···················· | ···················· | ····················· | ···················· | ···················· | ···················· |
| O | 5094 | hCoV-19/Gibraltar/204421212/2020\|EPI_ISL_613673\|20 | ···················· | ···················· | ····················· | ···················· | ······T············· | ···················· |
| O | 5095 | hCoV-19/Gibraltar/204421213/2020\|EPI_ISL_613674\|20 | ···················· | ···················· | ····················· | ···················· | ···················· | ···················· |
| O | 5096 | hCoV-19/Gibraltar/204421214/2020\|EPI_ISL_613675\|20 | ···················· | ···················· | ····················· | ···················· | ···················· | ···················· |
| O | 5097 | hCoV-19/Gibraltar/204421215/2020\|EPI_ISL_613676\|20 | ···················· | ···················· | ····················· | ···················· | ···················· | ···················· |
| O | 5098 | hCoV-19/Gibraltar/204421216/2020\|EPI_ISL_613677\|20 | ···················· | ···················· | ····················· | ···················· | ···················· | ···················· |
| O | 5099 | hCoV-19/Gibraltar/204421217/2020\|EPI_ISL_613678\|20 | ···················· | ···················· | ····················· | ···················· | ···················· | ···················· |
| O | 5100 | hCoV-19/Gibraltar/204421218/2020\|EPI_ISL_613679\|20 | ···················· | ···················· | ····················· | ···················· | ···················· | ···················· |
| O | 5101 | hCoV-19/Gibraltar/204421219/2020\|EPI_ISL_613680\|20 | ···················· | ···················· | ····················· | ···················· | ···················· | ···················· |
| O | 5102 | hCoV-19/Gibraltar/204421220/2020\|EPI_ISL_613681\|20 | ···················· | ···················· | ····················· | ···················· | ···················· | ···················· |
| O | 5103 | hCoV-19/Gibraltar/204421221/2020\|EPI_ISL_613682\|20 | ···················· | ···················· | ····················· | ···················· | ···················· | ···················· |
| O | 5104 | hCoV-19/Gibraltar/204421222/2020\|EPI_ISL_613683\|20 | ···················· | ·A·················· | ····················· | ···················· | ···················· | ··A················· |
| O | 5105 | hCoV-19/Gibraltar/204421224/2020\|EPI_ISL_613684\|20 | ···················· | ···················· | ····················· | ···················· | ···················· | ···················· |
| O | 5106 | hCoV-19/Gibraltar/204421225/2020\|EPI_ISL_613685\|20 | ···················· | ···················· | ····················· | ···················· | ···················· | ···················· |
| O | 5107 | hCoV-19/Gibraltar/204421226/2020\|EPI_ISL_613686\|20 | ···················· | ···················· | ····················· | ···················· | ···················· | ···················· |
| O | 5108 | hCoV-19/Gibraltar/204421227/2020\|EPI_ISL_613687\|20 | ···················· | ···················· | ····················· | ···················· | ···················· | ···················· |
| O | 5109 | hCoV-19/Gibraltar/204421228/2020\|EPI_ISL_613688\|20 | ···················· | ···················· | ····················· | ···················· | ···················· | ···················· |

| | | | | | | | | |
|---|---|---|---|---|---|---|---|---|
| O | 5110 | hCoV-19/Gibraltar/204421229/2020\|EPI_ISL_613689\|20 | ···················· | ···················· | ···················· | ···················· | ···················· | ···················· |
| O | 5111 | hCoV-19/Gibraltar/204421230/2020\|EPI_ISL_613690\|20 | ···················· | ·A·················· | ···················· | ···················· | ···················· | ···A················ |
| O | 5112 | hCoV-19/Gibraltar/204421231/2020\|EPI_ISL_613691\|20 | ···················· | ···················· | ···················· | ···················· | ···················· | ···················· |
| O | 5113 | hCoV-19/Gibraltar/204421232/2020\|EPI_ISL_613692\|20 | ···················· | ···················· | ···················· | ···················· | ···················· | ···················· |
| O | 5114 | hCoV-19/Gibraltar/204421233/2020\|EPI_ISL_613693\|20 | ···················· | ···················· | ···················· | ···················· | ···················· | ···················· |
| O | 5115 | hCoV-19/Gibraltar/204421234/2020\|EPI_ISL_613694\|20 | ···················· | ···················· | ···················· | ···················· | ···················· | ·····M·············· |
| O | 5116 | hCoV-19/Gibraltar/204421235/2020\|EPI_ISL_613695\|20 | ···················· | ···················· | ···················· | ···················· | ······T············· | ···················· |
| O | 5117 | hCoV-19/Gibraltar/204421237/2020\|EPI_ISL_613696\|20 | ···················· | ···················· | ···················· | ···················· | ···················· | ···················· |
| O | 5118 | hCoV-19/Gibraltar/204421238/2020\|EPI_ISL_613697\|20 | ···················· | ···················· | ···················· | ···················· | ···················· | ···················· |
| O | 5119 | hCoV-19/Gibraltar/204421239/2020\|EPI_ISL_613698\|20 | ···················· | ···················· | ···················· | ···················· | ···················· | ···················· |
| O | 5120 | hCoV-19/Gibraltar/204421240/2020\|EPI_ISL_613699\|20 | ···················· | ···················· | ···················· | ···················· | ···················· | ···················· |
| O | 5121 | hCoV-19/England/MILK-A7DACA/2020\|EPI_ISL_606460\|20 | ···················· | ···················· | ···················· | ···················· | ···················· | ···················· |
| O | 5122 | hCoV-19/England/MILK-A7C348/2020\|EPI_ISL_606461\|20 | ···················· | ···················· | ···················· | ···················· | ···················· | ···················· |
| O | 5123 | hCoV-19/England/MILK-A7C357/2020\|EPI_ISL_606463\|20 | ···················· | ···················· | ···················· | ···················· | ···················· | ···················· |
| O | 5124 | hCoV-19/England/MILK-A7C0E7/2020\|EPI_ISL_606465\|20 | ···················· | ···················· | ···················· | ···················· | ······T············· | ···················· |
| O | 5125 | hCoV-19/England/MILK-A7AF28/2020\|EPI_ISL_606466\|20 | ···················· | ···················· | ···················· | ···················· | ···················· | ···················· |
| O | 5126 | hCoV-19/England/MILK-A7C366/2020\|EPI_ISL_606467\|20 | ···················· | ···················· | ···················· | ···················· | ···················· | ···················· |
| O | 5127 | hCoV-19/England/MILK-A7DF43/2020\|EPI_ISL_606468\|20 | ···················· | ···················· | ···················· | ···················· | ···················· | ···················· |
| O | 5128 | hCoV-19/England/MILK-A7C0F6/2020\|EPI_ISL_606469\|20 | ···················· | ···················· | ···················· | ···················· | ···················· | ···················· |
| O | 5129 | hCoV-19/England/MILK-A7C384/2020\|EPI_ISL_606470\|20 | ···················· | ···················· | ···················· | ···················· | ···················· | ···················· |
| O | 5130 | hCoV-19/England/MILK-A7AA09/2020\|EPI_ISL_606471\|20 | ···················· | ···················· | ···················· | ···················· | ···················· | ···················· |
| O | 5131 | hCoV-19/England/MILK-A7D22C/2020\|EPI_ISL_606472\|20 | ···················· | ·A·················· | ···················· | ···················· | ···················· | ···················· |
| O | 5132 | hCoV-19/England/MILK-A7C6E5/2020\|EPI_ISL_606473\|20 | ···················· | ···················· | ···················· | ···················· | ···················· | ···················· |
| O | 5133 | hCoV-19/England/MILK-A7D21D/2020\|EPI_ISL_606474\|20 | ···················· | ···················· | ···················· | ···················· | ······T············· | ···················· |
| O | 5134 | hCoV-19/England/MILK-A7CA8F/2020\|EPI_ISL_606475\|20 | ···················· | ···················· | ···················· | ···················· | ···················· | ···················· |
| O | 5135 | hCoV-19/England/MILK-A7D426/2020\|EPI_ISL_606476\|20 | ···················· | ···················· | ···················· | ···················· | ···················· | ···················· |
| O | 5136 | hCoV-19/England/MILK-A7D2D1/2020\|EPI_ISL_606477\|20 | ···················· | ···················· | ···················· | ···················· | ···················· | ···················· |
| O | 5137 | hCoV-19/England/MILK-A7D1B6/2020\|EPI_ISL_606478\|20 | ···················· | ···················· | ···················· | ···················· | ···················· | ···················· |
| O | 5138 | hCoV-19/England/MILK-A7ADC4/2020\|EPI_ISL_606479\|20 | ···················· | ···················· | ···················· | ···················· | ···················· | ···················· |
| O | 5139 | hCoV-19/England/MILK-A7D3B0/2020\|EPI_ISL_606480\|20 | ···················· | ···················· | ···················· | ···················· | ···················· | ···················· |
| O | 5140 | hCoV-19/England/MILK-A7C427/2020\|EPI_ISL_606481\|20 | ···················· | ···················· | ···················· | ···················· | ···················· | ···················· |
| O | 5141 | hCoV-19/England/MILK-A7DEA0/2020\|EPI_ISL_606482\|20 | ···················· | ···················· | ···················· | ···················· | ···················· | ···················· |
| O | 5142 | hCoV-19/England/MILK-A7D699/2020\|EPI_ISL_606483\|20 | ···················· | ···················· | ···················· | ···················· | ···················· | ···················· |
| O | 5143 | hCoV-19/England/MILK-A7D6B7/2020\|EPI_ISL_606484\|20 | ···················· | ···················· | ···················· | ···················· | ···················· | ···················· |
| O | 5144 | hCoV-19/England/MILK-A7AEC1/2020\|EPI_ISL_606487\|20 | ···················· | ···················· | ···················· | ···················· | ···················· | ···················· |
| O | 5145 | hCoV-19/England/MILK-A7CB8C/2020\|EPI_ISL_606489\|20 | ···················· | ···················· | ···················· | ···················· | ······T············· | ···················· |
| O | 5146 | hCoV-19/England/MILK-A7C5D9/2020\|EPI_ISL_606490\|20 | ···················· | ···················· | ···················· | ···················· | ···················· | ···················· |
| O | 5147 | hCoV-19/England/MILK-A7C3C0/2020\|EPI_ISL_606491\|20 | ···················· | ···················· | ···················· | ···················· | ···················· | ···················· |
| O | 5148 | hCoV-19/England/MILK-A7A7D5/2020\|EPI_ISL_606492\|20 | ···················· | ···················· | ···················· | ···················· | ···················· | ···················· |
| O | 5149 | hCoV-19/England/MILK-A7AEA3/2020\|EPI_ISL_606493\|20 | ···················· | ···················· | ···················· | ···················· | ···················· | ···················· |
| O | 5150 | hCoV-19/England/MILK-A7D839/2020\|EPI_ISL_606494\|20 | ···················· | ···················· | ···················· | ···················· | ···················· | ···················· |
| O | 5151 | hCoV-19/England/MILK-A7CB31/2020\|EPI_ISL_606495\|20 | ···················· | ···················· | ···················· | ···················· | ···················· | ···················· |
| O | 5152 | hCoV-19/England/MILK-A7C621/2020\|EPI_ISL_606496\|20 | ···················· | ···················· | ···················· | ···················· | ···················· | ···················· |
| O | 5153 | hCoV-19/England/MILK-A7D71E/2020\|EPI_ISL_606497\|20 | ···················· | ···················· | ···················· | ···················· | ······T············· | ···················· |
| O | 5154 | hCoV-19/England/MILK-A7CD95/2020\|EPI_ISL_606498\|20 | ···················· | ···················· | ···················· | ···················· | ···················· | ···················· |
| O | 5155 | hCoV-19/England/MILK-A7DEFB/2020\|EPI_ISL_606499\|20 | ···················· | ···················· | ···················· | ···················· | ···················· | ···················· |
| O | 5156 | hCoV-19/England/MILK-A7D16B/2020\|EPI_ISL_606521\|20 | ···················· | ···················· | ···················· | ···················· | ···················· | ···················· |
| O | 5157 | hCoV-19/England/MILK-A7D769/2020\|EPI_ISL_606530\|20 | ···················· | ···················· | ···················· | ···················· | ···················· | ···················· |
| O | 5158 | hCoV-19/England/MILK-A7DC5B/2020\|EPI_ISL_606531\|20 | ···················· | ···················· | ···················· | ···················· | ···················· | ···················· |
| O | 5159 | hCoV-19/England/MILK-A7D611/2020\|EPI_ISL_606532\|20 | ···················· | ···················· | ···················· | ···················· | ···················· | ···················· |
| O | 5160 | hCoV-19/England/MILK-A7D4DB/2020\|EPI_ISL_606533\|20 | ···················· | ···················· | ···················· | ···················· | ···················· | ···················· |
| O | 5161 | hCoV-19/England/MILK-A7DCE2/2020\|EPI_ISL_606534\|20 | ···················· | ···················· | ···················· | ···················· | ···················· | ···················· |
| O | 5162 | hCoV-19/England/MILK-A7AEB2/2020\|EPI_ISL_606535\|20 | ···················· | ···················· | ···················· | ···················· | ···················· | ···················· |
| O | 5163 | hCoV-19/England/MILK-A7C375/2020\|EPI_ISL_606536\|20 | ···················· | ···················· | ···················· | ···················· | ···················· | ···················· |
| O | 5164 | hCoV-19/England/MILK-A7CB40/2020\|EPI_ISL_606537\|20 | ···················· | ···················· | ···················· | ···················· | ···················· | ···················· |
| O | 5165 | hCoV-19/England/MILK-A7D57E/2020\|EPI_ISL_606538\|20 | ···················· | ···················· | ···················· | ···················· | ···················· | ···················· |
| O | 5166 | hCoV-19/England/MILK-A7DDD0/2020\|EPI_ISL_606539\|20 | ···················· | ···················· | ···················· | ···················· | ···················· | ···················· |
| O | 5167 | hCoV-19/England/ALDP-A703DB/2020\|EPI_ISL_607700\|20 | ···················· | ···················· | ···················· | ···················· | ···················· | ···················· |
| O | 5168 | hCoV-19/England/ALDP-A6EFCD/2020\|EPI_ISL_607701\|20 | ···················· | ···················· | ···················· | ···················· | ···················· | ···················· |
| O | 5169 | hCoV-19/England/ALDP-A6E859/2020\|EPI_ISL_607702\|20 | ···················· | ···················· | ···················· | ···················· | ···················· | ···················· |
| O | 5170 | hCoV-19/England/ALDP-A6F481/2020\|EPI_ISL_607703\|20 | ···················· | ···················· | ···················· | ···················· | ···················· | ···················· |
| O | 5171 | hCoV-19/England/ALDP-A6F3DF/2020\|EPI_ISL_607704\|20 | ···················· | ···················· | ···················· | ···················· | ···················· | ···················· |
| O | 5172 | hCoV-19/England/ALDP-A6ED0F/2020\|EPI_ISL_607705\|20 | ···················· | ···················· | ···················· | ···················· | ···················· | ···················· |
| O | 5173 | hCoV-19/England/ALDP-A6E7C5/2020\|EPI_ISL_607706\|20 | ···················· | ···················· | ···················· | ···················· | ···················· | ···················· |
| O | 5174 | hCoV-19/England/ALDP-A70353/2020\|EPI_ISL_607707\|20 | ···················· | ·A·················· | ···················· | ···················· | ···················· | ···················· |
| O | 5175 | hCoV-19/England/ALDP-A70748/2020\|EPI_ISL_607790\|20 | ···················· | ···················· | ···················· | ···················· | ···················· | ···················· |
| O | 5176 | hCoV-19/England/ALDP-A6DEB2/2020\|EPI_ISL_607791\|20 | ···················· | ···················· | ···················· | ···················· | ···················· | ···················· |
| O | 5177 | hCoV-19/England/ALDP-A702ED/2020\|EPI_ISL_607792\|20 | ···················· | ···················· | ···················· | ···················· | ···················· | ···················· |
| O | 5178 | hCoV-19/England/ALDP-A6E83B/2020\|EPI_ISL_607793\|20 | ···················· | ···················· | ···················· | ···················· | ···················· | ···················· |
| O | 5179 | hCoV-19/England/ALDP-A6E17C/2020\|EPI_ISL_607794\|20 | ···················· | ···················· | ···················· | ···················· | ···················· | ···················· |
| O | 5180 | hCoV-19/England/ALDP-A702DE/2020\|EPI_ISL_607795\|20 | ···················· | ···················· | ···················· | ···················· | ···················· | ···················· |
| O | 5181 | hCoV-19/England/ALDP-A7061E/2020\|EPI_ISL_607796\|20 | ···················· | ···················· | ···················· | ···················· | ···················· | ···················· |
| O | 5182 | hCoV-19/England/ALDP-A6F64F/2020\|EPI_ISL_607797\|20 | ···················· | ···················· | ···················· | ···················· | ···················· | ···················· |
| O | 5183 | hCoV-19/England/ALDP-A6E992/2020\|EPI_ISL_607798\|20 | ···················· | ···················· | ···················· | ···················· | ···················· | ···················· |
| O | 5184 | hCoV-19/England/ALDP-A6FA16/2020\|EPI_ISL_607799\|20 | ···················· | ···················· | ···················· | ···················· | ···················· | ···················· |
| O | 5185 | hCoV-19/England/ALDP-A6D90C/2020\|EPI_ISL_607851\|20 | ···················· | ···················· | ···················· | ···················· | ···················· | ···················· |
| O | 5186 | hCoV-19/England/ALDP-A6F4BE/2020\|EPI_ISL_607861\|20 | ···················· | ···················· | ···················· | ···················· | ···················· | ···················· |
| O | 5187 | hCoV-19/England/ALDP-A6F612/2020\|EPI_ISL_607862\|20 | ···················· | ···················· | ···················· | ···················· | ···················· | ···················· |
| O | 5188 | hCoV-19/England/ALDP-A6D8A5/2020\|EPI_ISL_607863\|20 | ···················· | ···················· | ···················· | ···················· | ···················· | ···················· |
| O | 5189 | hCoV-19/England/ALDP-A6ECF3/2020\|EPI_ISL_607864\|20 | ···················· | ···················· | ···················· | ···················· | ···················· | ···················· |

O 5190 hCoV-19/England/ALDP-A6E25B/2020|EPI_ISL_607865|20 ···················· ···················· ···················· ···················· ···················· ····················
O 5191 hCoV-19/England/ALDP-A6DF82/2020|EPI_ISL_607868|20 ···················· ···················· ···················· ···················· ···················· ····················
O 5192 hCoV-19/England/ALDP-A703BD/2020|EPI_ISL_607869|20 ···················· ···················· ···················· ···················· ···················· ····················
O 5193 hCoV-19/England/ALDP-A92889/2020|EPI_ISL_611000|20 ···················· ···················· ···················· ···················· ···················· ····················
O 5194 hCoV-19/England/ALDP-A93FB0/2020|EPI_ISL_611002|20 ···················· ···················· ···················· ···················· ···················· ····················
O 5195 hCoV-19/England/ALDP-A93E1D/2020|EPI_ISL_611003|20 ···················· ···················· ···················· ···················· ···················· ····················
O 5196 hCoV-19/England/ALDP-A93712/2020|EPI_ISL_611004|20 ···················· ···················· ···················· ···················· ···················· ····················
O 5197 hCoV-19/England/ALDP-A92E78/2020|EPI_ISL_611005|20 ···················· ···················· ···················· ···················· ···················· ····················
O 5198 hCoV-19/England/ALDP-A93E68/2020|EPI_ISL_611007|20 ···················· ···················· ···················· ···················· ···················· ····················
O 5199 hCoV-19/England/BIRM-65CD8/2020|EPI_ISL_612220|202 ···················· ···················· ···················· ···················· ···················· ····················
O 5201 hCoV-19/England/BIRM-65D3F/2020|EPI_ISL_612222|202 ···················· ·A·················· ···················· ···················· ···················· ··A·················
O 5202 hCoV-19/England/BIRM-65D4E/2020|EPI_ISL_612223|202 ···················· ···················· ···················· ···················· ···················· ····················
O 5203 hCoV-19/England/BIRM-65D5D/2020|EPI_ISL_612224|202 ···················· ·A·················· ···················· ···················· ···················· ··A·················
O 5204 hCoV-19/England/BIRM-65D8A/2020|EPI_ISL_612225|202 ···················· ···················· ···················· ···················· ···················· ····················
O 5205 hCoV-19/England/BIRM-65D99/2020|EPI_ISL_612226|202 ···················· ·A·················· ···················· ···················· ···················· ··A·················
O 5206 hCoV-19/England/BIRM-65DA8/2020|EPI_ISL_612227|202 ···················· ···················· ···················· ···················· ···················· ····················
O 5208 hCoV-19/England/BIRM-66396/2020|EPI_ISL_612260|202 ···················· ·A·················· ···················· ···················· ···················· ··A·················
O 5209 hCoV-19/England/CAMB-1B6624/2020|EPI_ISL_612261|20 ···················· ···················· ···················· ···················· ···················· ····················
O 5210 hCoV-19/England/CAMB-1B6642/2020|EPI_ISL_612262|20 ···················· ···················· ···················· ···················· ···················· ····················
O 5211 hCoV-19/England/CAMB-1B6660/2020|EPI_ISL_612263|20 ···················· ···················· ···················· ···················· ···················· ····················
O 5212 hCoV-19/England/CAMB-1B668E/2020|EPI_ISL_612264|20 ···················· ···················· ···················· ···················· ···················· ····················
O 5213 hCoV-19/England/CAMB-1B6A37/2020|EPI_ISL_612265|20 ···················· ···················· ···················· ···················· ···················· ····················
O 5215 hCoV-19/England/CAMB-1B6A64/2020|EPI_ISL_612267|20 ···················· ···················· ···················· ···················· ···················· ····················
O 5216 hCoV-19/England/CAMB-1B6A73/2020|EPI_ISL_612268|20 ···················· ···················· ···················· ···················· ···················· ····················
O 5217 hCoV-19/England/CAMB-1B6A82/2020|EPI_ISL_612269|20 ···················· ···················· ···················· ···················· ···················· ····················
O 5218 hCoV-19/England/CAMB-1B6AA0/2020|EPI_ISL_612270|20 ···················· ···················· ···················· ···················· ···················· ····················
O 5219 hCoV-19/England/CAMB-1B6ABF/2020|EPI_ISL_612271|20 ···················· ···················· ···················· ···················· ···················· ····················
O 5220 hCoV-19/England/CAMB-1B6ACE/2020|EPI_ISL_612272|20 ···················· ···················· ···················· ···················· ···················· ····················
O 5221 hCoV-19/England/CAMB-1B6B43/2020|EPI_ISL_612273|20 ···················· ···················· ···················· ···················· ···················· ····················
O 5222 hCoV-19/England/CAMB-1B6C04/2020|EPI_ISL_612274|20 ···················· ···················· ···················· ···················· ···················· ····················
O 5223 hCoV-19/England/CAMB-1B6C13/2020|EPI_ISL_612275|20 ···················· ···················· ···················· ···················· ···················· ····················
O 5224 hCoV-19/England/CAMB-1B6C6E/2020|EPI_ISL_612276|20 ···················· ···················· ···················· ···················· ···················· ····················
O 5225 hCoV-19/England/CAMB-1B6CAA/2020|EPI_ISL_612277|20 ···················· ···················· ···················· ···················· ···················· ····················
O 5226 hCoV-19/England/CAMB-1B6D01/2020|EPI_ISL_612278|20 ···················· ···················· ···················· ···················· ···················· ····················
O 5227 hCoV-19/England/CAMB-1B6D3E/2020|EPI_ISL_612279|20 ···················· ···················· ···················· ···················· ···················· ····················
O 5228 hCoV-19/England/CAMB-1B6D5C/2020|EPI_ISL_612280|20 ···················· ···················· ···················· ···················· ···················· ····················
O 5229 hCoV-19/England/CAMB-1B6DB6/2020|EPI_ISL_612281|20 ···················· ···················· ···················· ···················· ······T············· ····················
O 5230 hCoV-19/England/CAMB-1B6DF2/2020|EPI_ISL_612282|20 ···················· ···················· ···················· ···················· ···················· ····················
O 5231 hCoV-19/England/CAMB-1B6E59/2020|EPI_ISL_612283|20 ···················· ···················· ···················· ···················· ···················· ····················
O 5232 hCoV-19/Scotland/CVR4630/2020|EPI_ISL_612284|2020- ···················· ···················· ···················· ···················· ···················· ····················
O 5233 hCoV-19/Scotland/CVR4632/2020|EPI_ISL_612285|2020- ···················· ···················· ···················· ···················· ···················· ····················
O 5234 hCoV-19/Scotland/CVR4633/2020|EPI_ISL_612286|2020- ···················· ···················· ···················· ···················· ···················· ····················
O 5235 hCoV-19/Scotland/CVR4634/2020|EPI_ISL_612287|2020- ···················· ···················· ···················· ···················· ···················· ····················
O 5236 hCoV-19/Scotland/CVR4641/2020|EPI_ISL_612288|2020- ···················· ···················· ···················· ···················· ···················· ····················
O 5237 hCoV-19/Scotland/CVR4643/2020|EPI_ISL_612289|2020- ···················· ···················· ···················· ···················· ···················· NN·N················ 1
O 5238 hCoV-19/Scotland/CVR4645/2020|EPI_ISL_612290|2020- ···················· ···················· ···················· ···················· ···················· ····················
O 5239 hCoV-19/Scotland/CVR4646/2020|EPI_ISL_612291|2020- ···················· ···················· ···················· ···················· ···················· ····················
O 5240 hCoV-19/Scotland/CVR4647/2020|EPI_ISL_612292|2020- ···················· ···················· ···················· ···················· ···················· ····················
O 5241 hCoV-19/Scotland/CVR4648/2020|EPI_ISL_612293|2020- ···················· ···················· ···················· ···················· ···················· NN·············N···· 1
O 5242 hCoV-19/Scotland/CVR4652/2020|EPI_ISL_612294|2020- ···················· ···················· ···················· ···················· ···················· ····················
O 5243 hCoV-19/Scotland/CVR4653/2020|EPI_ISL_612295|2020- ···················· ···················· ···················· ···················· ···················· ····················
O 5244 hCoV-19/Scotland/CVR4657/2020|EPI_ISL_612296|2020- ···················· ···················· ···················· ···················· ···················· ····················
O 5245 hCoV-19/Scotland/CVR4659/2020|EPI_ISL_612297|2020- ···················· ···················· ···················· ···················· ···················· ····················
O 5246 hCoV-19/Scotland/CVR4660/2020|EPI_ISL_612298|2020- ···················· ···················· ···················· ···················· ··············N····· N···················
O 5247 hCoV-19/Scotland/CVR4661/2020|EPI_ISL_612299|2020- ···················· ···················· ···················· ···················· ···················· ····················
O 5248 hCoV-19/Scotland/CVR4662/2020|EPI_ISL_612300|2020- ···················· ···················· ···················· ···················· ···················· ····················
O 5249 hCoV-19/Scotland/CVR4664/2020|EPI_ISL_612301|2020- ···················· ···················· ···················· ···················· ···················· ····················
O 5250 hCoV-19/Scotland/CVR4665/2020|EPI_ISL_612302|2020- ···················· ···················· ···················· ···················· ···················· NN··················
O 5251 hCoV-19/Scotland/CVR4666/2020|EPI_ISL_612303|2020- ···················· ···················· ···················· ···················· ···················· ····················
O 5253 hCoV-19/Scotland/CVR4669/2020|EPI_ISL_612305|2020- ···················· ···················· ···················· ···················· ···················· ···N···NN··········· 1
O 5254 hCoV-19/Scotland/CVR4677/2020|EPI_ISL_612306|2020- ···················· ···················· ···················· ···················· ···················· ····················
O 5255 hCoV-19/Scotland/CVR4678/2020|EPI_ISL_612307|2020- ···················· ···················· ···················· ···················· ···················· ····················
O 5256 hCoV-19/Scotland/CVR4680/2020|EPI_ISL_612308|2020- ···················· ···················· ···················· ···················· ···················· ····················
O 5257 hCoV-19/Scotland/CVR4681/2020|EPI_ISL_612309|2020- ···················· ···················· ···················· ···················· ···················· ····················
O 5258 hCoV-19/Scotland/CVR4688/2020|EPI_ISL_612310|2020- ···················· ···················· ···················· ···················· ···················· ····················
O 5259 hCoV-19/Scotland/CVR4689/2020|EPI_ISL_612311|2020- ···················· ···················· ···················· ···················· ···················· ····················
O 5260 hCoV-19/Scotland/CVR4690/2020|EPI_ISL_612312|2020- ···················· ···················· ···················· ···················· ···················· ····················
O 5262 hCoV-19/Scotland/EDB8274/2020|EPI_ISL_612314|2020- ···················· ···················· ···················· ···················· ···················· ····················
O 5263 hCoV-19/Scotland/EDB8275/2020|EPI_ISL_612315|2020- ···················· ···················· ···················· ···················· ···················· ····················
O 5264 hCoV-19/Scotland/EDB8295/2020|EPI_ISL_612316|2020- ···················· ···················· ···················· ···················· ···················· ····················
O 5265 hCoV-19/Scotland/EDB8298/2020|EPI_ISL_612317|2020- ···················· ···················· ···················· ···················· ···················· ····················
O 5266 hCoV-19/Scotland/EDB8299/2020|EPI_ISL_612318|2020- ···················· ···················· ···················· ···················· ···················· ····················
O 5267 hCoV-19/Scotland/EDB8300/2020|EPI_ISL_612319|2020- ···················· ···················· ···················· ···················· ···················· ····················
O 5268 hCoV-19/Scotland/EDB8326/2020|EPI_ISL_612320|2020- ···················· ···················· ···················· ···················· ···················· ····················
O 5269 hCoV-19/Scotland/EDB8327/2020|EPI_ISL_612321|2020- ···················· ···················· ···················· ···················· ···················· ····················
O 5270 hCoV-19/Scotland/EDB8328/2020|EPI_ISL_612322|2020- ···················· ·A·················· ···················· ···················· ···················· ··A·················
O 5271 hCoV-19/Scotland/EDB8329/2020|EPI_ISL_612323|2020- ···················· ···················· ···················· ···················· ···················· ····················
O 5272 hCoV-19/Scotland/EDB8330/2020|EPI_ISL_612324|2020- ···················· ···················· ···················· ···················· ···················· ····················
O 5273 hCoV-19/Scotland/EDB8332/2020|EPI_ISL_612325|2020- ···················· ···················· ···················· ···················· ···················· ····················
O 5274 hCoV-19/Scotland/EDB8334/2020|EPI_ISL_612326|2020- ···················· ···················· ···················· ···················· ···················· ····················

| | | | | | | | |
|---|---|---|---|---|---|---|---|
| O | 5275 hCoV-19/Scotland/EDB8335/2020\|EPI_ISL_612327\|2020- | ···················· | ···················· | ···················· | ···················· | ···················· | ···················· |
| O | 5276 hCoV-19/Scotland/EDB8340/2020\|EPI_ISL_612328\|2020- | ···················· | ···················· | ···················· | ···················· | ······T············· | ···················· |
| O | 5277 hCoV-19/Scotland/EDB8341/2020\|EPI_ISL_612329\|2020- | ···················· | ···················· | ···················· | ···················· | ···················· | ···················· |
| O | 5278 hCoV-19/Scotland/EDB8342/2020\|EPI_ISL_612330\|2020- | ···················· | ···················· | ···················· | ···················· | ······T············· | ···················· |
| O | 5279 hCoV-19/Scotland/EDB8343/2020\|EPI_ISL_612331\|2020- | ···················· | ···················· | ···················· | ···················· | ···················· | ···················· |
| O | 5280 hCoV-19/Scotland/EDB8344/2020\|EPI_ISL_612332\|2020- | ···················· | ···················· | ···················· | ···················· | ······T············· | ···················· |
| O | 5281 hCoV-19/Scotland/EDB8345/2020\|EPI_ISL_612333\|2020- | ···················· | ···················· | ···················· | ···················· | ···················· | ···················· |
| O | 5282 hCoV-19/Scotland/EDB8346/2020\|EPI_ISL_612334\|2020- | ···················· | ···················· | ···················· | ···················· | ···················· | ···················· |
| O | 5283 hCoV-19/Scotland/EDB8347/2020\|EPI_ISL_612335\|2020- | ···················· | ···················· | ···················· | ···················· | ···················· | ···················· |
| O | 5284 hCoV-19/Scotland/EDB8348/2020\|EPI_ISL_612336\|2020- | ···················· | ···················· | ···················· | ···················· | ···················· | ···················· |
| O | 5285 hCoV-19/Scotland/EDB8349/2020\|EPI_ISL_612337\|2020- | ···················· | ···················· | ···················· | ···················· | ···················· | ···················· |
| O | 5286 hCoV-19/Scotland/EDB8350/2020\|EPI_ISL_612338\|2020- | ···················· | ···················· | ···················· | ···················· | ···················· | ···················· |
| O | 5287 hCoV-19/France/BRE-BR9061/2020\|EPI_ISL_613550\|2020 | ···················· | ···················· | ···················· | ···················· | ···················· | ···················· |
| O | 5288 hCoV-19/France/BRE-BR9062/2020\|EPI_ISL_613551\|2020 | ···················· | ···················· | ···················· | ···················· | ···················· | ···················· |
| O | 5289 hCoV-19/France/BRE-BR9063/2020\|EPI_ISL_613552\|2020 | ···················· | ···················· | ···················· | ···················· | ···················· | ···················· |
| O | 5290 hCoV-19/France/BRE-BR9064/2020\|EPI_ISL_613553\|2020 | ···················· | ···················· | ···················· | ···················· | ···················· | ···················· |
| O | 5291 hCoV-19/France/BRE-BR9065/2020\|EPI_ISL_613554\|2020 | ···················· | ···················· | ···················· | ···················· | ···················· | ···················· |
| O | 5292 hCoV-19/France/BRE-BR9066/2020\|EPI_ISL_613555\|2020 | ···················· | ···················· | ···················· | ···················· | ···················· | ···················· |
| O | 5293 hCoV-19/France/BRE-BR9067/2020\|EPI_ISL_613556\|2020 | ···················· | ···················· | ···················· | ···················· | ···················· | ···················· |
| O | 5294 hCoV-19/France/BRE-BR9068/2020\|EPI_ISL_613557\|2020 | ···················· | ···················· | ···················· | ···················· | ···················· | ···················· |
| O | 5295 hCoV-19/France/BRE-BR9070/2020\|EPI_ISL_613558\|2020 | ···················· | ···················· | ···················· | ···················· | ···················· | ···················· |
| O | 5296 hCoV-19/France/BRE-BR9071/2020\|EPI_ISL_613559\|2020 | ···················· | ···················· | ···················· | ···················· | ···················· | ···············A···· |
| O | 5297 hCoV-19/England/204262693/2020\|EPI_ISL_613590\|2020 | ···················· | ···················· | ···················· | ···················· | ···················· | ···················· |
| O | 5298 hCoV-19/England/204280114/2020\|EPI_ISL_613591\|2020 | ···················· | ···················· | ···················· | ···················· | ···················· | ···················· |
| O | 5299 hCoV-19/England/204280131/2020\|EPI_ISL_613592\|2020 | ···················· | ···················· | ···················· | ···················· | ···················· | ···················· |
| O | 5300 hCoV-19/England/204280140/2020\|EPI_ISL_613593\|2020 | ···················· | ···················· | ···················· | ···················· | ···················· | ···················· |
| O | 5301 hCoV-19/England/204280143/2020\|EPI_ISL_613594\|2020 | ···················· | ···················· | ···················· | ···················· | ···················· | ···················· |
| O | 5302 hCoV-19/England/204280144/2020\|EPI_ISL_613595\|2020 | ···················· | ···················· | ···················· | ···················· | ···················· | ···················· |
| O | 5303 hCoV-19/England/204280154/2020\|EPI_ISL_613596\|2020 | ···················· | ···················· | ···················· | ···················· | ···················· | ···················· |
| O | 5304 hCoV-19/England/204280156/2020\|EPI_ISL_613597\|2020 | ···················· | ···················· | ···················· | ···················· | ···················· | ···················· |
| O | 5305 hCoV-19/England/204281018/2020\|EPI_ISL_613598\|2020 | ···················· | ···················· | ···················· | ···················· | ···················· | ···················· |
| O | 5306 hCoV-19/England/204281813/2020\|EPI_ISL_613599\|2020 | ···················· | ···················· | ···················· | ···················· | ···················· | ···················· |
| O | 5307 hCoV-19/England/204281817/2020\|EPI_ISL_613600\|2020 | ···················· | ···················· | ···················· | ···················· | ···················· | ···················· |
| O | 5308 hCoV-19/England/204290227/2020\|EPI_ISL_613601\|2020 | ···················· | ···················· | ···················· | ···················· | ···················· | ···················· |
| O | 5309 hCoV-19/England/204290228/2020\|EPI_ISL_613602\|2020 | ···················· | ···················· | ···················· | ···················· | ···················· | ···················· |
| O | 5310 hCoV-19/England/204290229/2020\|EPI_ISL_613603\|2020 | ···················· | ···················· | ···················· | ···················· | ···················· | ···················· |
| O | 5311 hCoV-19/England/204290230/2020\|EPI_ISL_613604\|2020 | ···················· | ···················· | ···················· | ···················· | ···················· | ···················· |
| O | 5312 hCoV-19/England/204290231/2020\|EPI_ISL_613605\|2020 | ···················· | ···················· | ···················· | ···················· | ···················· | ···················· |
| O | 5313 hCoV-19/England/204290232/2020\|EPI_ISL_613606\|2020 | ···················· | ···················· | ···················· | ···················· | ···················· | ···················· |
| O | 5314 hCoV-19/England/204290233/2020\|EPI_ISL_613607\|2020 | ···················· | ···················· | ···················· | ···················· | ···················· | ···················· |
| O | 5315 hCoV-19/England/204290235/2020\|EPI_ISL_613608\|2020 | ···················· | ···················· | ···················· | ···················· | ···················· | ···················· |
| O | 5316 hCoV-19/England/204290236/2020\|EPI_ISL_613609\|2020 | ···················· | ···················· | ···················· | ···················· | ···················· | ···················· |
| O | 5317 hCoV-19/England/204290237/2020\|EPI_ISL_613610\|2020 | ···················· | ···················· | ···················· | ···················· | ···················· | ···················· |
| O | 5318 hCoV-19/England/204290338/2020\|EPI_ISL_613611\|2020 | ···················· | ···················· | ···················· | ···················· | ···················· | ···················· |
| O | 5319 hCoV-19/England/204290511/2020\|EPI_ISL_613612\|2020 | ···················· | ···················· | ···················· | ···················· | ···················· | ···················· |
| O | 5320 hCoV-19/England/204290516/2020\|EPI_ISL_613613\|2020 | ···················· | ···················· | ···················· | ···················· | ···················· | ···················· |
| O | 5321 hCoV-19/England/204290519/2020\|EPI_ISL_613614\|2020 | ···················· | ···················· | ···················· | ···················· | ···················· | ···················· |
| O | 5322 hCoV-19/England/204291000/2020\|EPI_ISL_613615\|2020 | ···················· | ···················· | ···················· | ···················· | ···················· | ···················· |
| O | 5323 hCoV-19/England/204291001/2020\|EPI_ISL_613616\|2020 | ···················· | ···················· | ···················· | ···················· | ···················· | ···················· |
| O | 5324 hCoV-19/England/204291002/2020\|EPI_ISL_613617\|2020 | ···················· | ···················· | ···················· | ···················· | ···················· | ···················· |
| O | 5325 hCoV-19/England/204291004/2020\|EPI_ISL_613618\|2020 | ···················· | ···················· | ···················· | ···················· | ···················· | ···················· |
| O | 5326 hCoV-19/England/204291008/2020\|EPI_ISL_613619\|2020 | ···················· | ···················· | ···················· | ···················· | ···················· | ···················· |
| O | 5327 hCoV-19/England/204291009/2020\|EPI_ISL_613620\|2020 | ···················· | ···················· | ···················· | ···················· | ···················· | ···················· |
| O | 5328 hCoV-19/England/204291010/2020\|EPI_ISL_613621\|2020 | ···················· | ···················· | ···················· | ···················· | ···················· | ···················· |
| O | 5329 hCoV-19/England/204291011/2020\|EPI_ISL_613622\|2020 | ···················· | ···················· | ···················· | ···················· | ···················· | ···················· |
| O | 5330 hCoV-19/England/204291034/2020\|EPI_ISL_613623\|2020 | ···················· | ···················· | ···················· | ···················· | ···················· | ···················· |
| O | 5331 hCoV-19/England/204291085/2020\|EPI_ISL_613624\|2020 | ···················· | ···················· | ···················· | ···················· | ···················· | ···················· |
| O | 5332 hCoV-19/England/204291136/2020\|EPI_ISL_613625\|2020 | ···················· | ···················· | ···················· | ···················· | ···················· | ···················· |
| O | 5333 hCoV-19/England/204291593/2020\|EPI_ISL_613626\|2020 | ···················· | ···················· | ···················· | ···················· | ···················· | ···················· |
| O | 5334 hCoV-19/England/204291681/2020\|EPI_ISL_613627\|2020 | ···················· | ···················· | ···················· | ···················· | ···················· | ···················· |
| O | 5335 hCoV-19/England/204291821/2020\|EPI_ISL_613628\|2020 | ···················· | ···················· | ···················· | ···················· | ···················· | ···················· |
| O | 5336 hCoV-19/England/204291822/2020\|EPI_ISL_613629\|2020 | ···················· | ···················· | ···················· | ···················· | ···················· | ···················· |
| O | 5337 hCoV-19/England/204291824/2020\|EPI_ISL_613630\|2020 | ···················· | ···················· | ···················· | ···················· | ···················· | ···················· |
| O | 5338 hCoV-19/England/204291975/2020\|EPI_ISL_613631\|2020 | ···················· | ···················· | ···················· | ···················· | ···················· | ···················· |
| O | 5339 hCoV-19/England/204291979/2020\|EPI_ISL_613632\|2020 | ···················· | ···················· | ···················· | ···················· | ···················· | ···················· |
| O | 5340 hCoV-19/England/204291982/2020\|EPI_ISL_613633\|2020 | ···················· | ···················· | ···················· | ···················· | ···················· | ···················· |
| O | 5341 hCoV-19/England/204400614/2020\|EPI_ISL_613634\|2020 | ···················· | ···················· | ···················· | ···················· | ···················· | ···················· |
| O | 5342 hCoV-19/England/204400615/2020\|EPI_ISL_613635\|2020 | ···················· | ···················· | ···················· | ···················· | ···················· | ···················· |
| O | 5343 hCoV-19/Gibraltar/204421172/2020\|EPI_ISL_613636\|20 | ···················· | ···················· | ···················· | ···················· | ···················· | ···················· |
| O | 5344 hCoV-19/Gibraltar/204421174/2020\|EPI_ISL_613637\|20 | ···················· | ···················· | ···················· | ···················· | ···················· | ···················· |
| O | 5345 hCoV-19/Gibraltar/204421175/2020\|EPI_ISL_613638\|20 | ···················· | ···················· | ···················· | ···················· | ···················· | ···················· |
| O | 5346 hCoV-19/Gibraltar/204421176/2020\|EPI_ISL_613639\|20 | ···················· | ···················· | ···················· | ···················· | ···················· | ···················· |
| O | 5347 hCoV-19/Gibraltar/204421177/2020\|EPI_ISL_613640\|20 | ···················· | ···················· | ···················· | ···················· | ···················· | ···················· |
| O | 5348 hCoV-19/Gibraltar/204421179/2020\|EPI_ISL_613641\|20 | ···················· | ···················· | ···················· | ···················· | ···················· | ···················· |
| O | 5349 hCoV-19/Gibraltar/204421180/2020\|EPI_ISL_613642\|20 | ···················· | ···················· | ···················· | ···················· | ···················· | ···················· |
| O | 5350 hCoV-19/Gibraltar/204421182/2020\|EPI_ISL_613643\|20 | ···················· | ···················· | ···················· | ···················· | ···················· | ···················· |
| O | 5351 hCoV-19/Gibraltar/204421183/2020\|EPI_ISL_613644\|20 | ···················· | ···················· | ···················· | ···················· | ···················· | ···················· |
| O | 5352 hCoV-19/Gibraltar/204421184/2020\|EPI_ISL_613645\|20 | ···················· | ···················· | ···················· | ···················· | ···················· | ···················· |
| O | 5353 hCoV-19/Gibraltar/204421185/2020\|EPI_ISL_613646\|20 | ···················· | ···················· | ···················· | ···················· | ···················· | ···················· |
| O | 5354 hCoV-19/Gibraltar/204421186/2020\|EPI_ISL_613647\|20 | ···················· | ···················· | ···················· | ···················· | ···················· | ···················· |

| | | | | | | | | |
|---|---|---|---|---|---|---|---|---|
| O | 5355 | hCoV-19/Gibraltar/204421187/2020\|EPI_ISL_613648\|20 | ···················· | ···················· | ····················· | ···················· | ···················· | ···················· |
| O | 5356 | hCoV-19/Gibraltar/204421188/2020\|EPI_ISL_613649\|20 | ···················· | ···················· | ····················· | ···················· | ···················· | ···················· |
| O | 5357 | hCoV-19/Gibraltar/204421189/2020\|EPI_ISL_613650\|20 | ···················· | ···················· | ····················· | ···················· | ···················· | ···················· |
| O | 5358 | hCoV-19/Gibraltar/204421190/2020\|EPI_ISL_613651\|20 | ···················· | ···················· | ····················· | ···················· | ···················· | ···················· |
| O | 5359 | hCoV-19/Gibraltar/204421191/2020\|EPI_ISL_613652\|20 | ···················· | ···················· | ····················· | ···················· | ······T············· | ···················· |
| O | 5360 | hCoV-19/Gibraltar/204421192/2020\|EPI_ISL_613653\|20 | ···················· | ···················· | ····················· | ···················· | ···················· | ···················· |
| O | 5361 | hCoV-19/Gibraltar/204421193/2020\|EPI_ISL_613654\|20 | ···················· | ···················· | ····················· | ···················· | ···················· | ···················· |
| O | 5362 | hCoV-19/Gibraltar/204421194/2020\|EPI_ISL_613655\|20 | ···················· | ···················· | ····················· | ···················· | ···················· | ···················· |
| O | 5363 | hCoV-19/Gibraltar/204421195/2020\|EPI_ISL_613656\|20 | ···················· | ···················· | ····················· | ···················· | ···················· | ···················· |
| O | 5364 | hCoV-19/Gibraltar/204421196/2020\|EPI_ISL_613657\|20 | ···················· | ···················· | ····················· | ···················· | ···················· | ···················· |
| O | 5365 | hCoV-19/Gibraltar/204421197/2020\|EPI_ISL_613658\|20 | ···················· | ···················· | ····················· | ···················· | ···················· | ···················· |
| O | 5366 | hCoV-19/Gibraltar/204421198/2020\|EPI_ISL_613659\|20 | ···················· | ···················· | ····················· | ···················· | ···················· | ···················· |
| O | 5367 | hCoV-19/Gibraltar/204421199/2020\|EPI_ISL_613660\|20 | ···················· | ···················· | ····················· | ···················· | ···················· | ···················· |
| O | 5368 | hCoV-19/Gibraltar/204421200/2020\|EPI_ISL_613661\|20 | ···················· | ···················· | ····················· | ···················· | ···················· | ···················· |
| O | 5369 | hCoV-19/Gibraltar/204421201/2020\|EPI_ISL_613662\|20 | ···················· | ···················· | ····················· | ···················· | ···················· | ···················· |
| O | 5370 | hCoV-19/Gibraltar/204421202/2020\|EPI_ISL_613663\|20 | ···················· | ···················· | ····················· | ···················· | ···················· | ···················· |
| O | 5371 | hCoV-19/Gibraltar/204421203/2020\|EPI_ISL_613664\|20 | ···················· | ···················· | ····················· | ···················· | ···················· | ···················· |
| O | 5372 | hCoV-19/Gibraltar/204421204/2020\|EPI_ISL_613665\|20 | ···················· | ···················· | ····················· | ···················· | ···················· | ···················· |
| O | 5373 | hCoV-19/Gibraltar/204421205/2020\|EPI_ISL_613666\|20 | ···················· | ···················· | ····················· | ···················· | ···················· | ···················· |
| O | 5374 | hCoV-19/Gibraltar/204421206/2020\|EPI_ISL_613667\|20 | ···················· | ···················· | ····················· | ···················· | ···················· | ···················· |
| O | 5375 | hCoV-19/Gibraltar/204421207/2020\|EPI_ISL_613668\|20 | ···················· | ···················· | ····················· | ···················· | ······T············· | ···················· |
| O | 5376 | hCoV-19/Gibraltar/204421208/2020\|EPI_ISL_613669\|20 | ···················· | ···················· | ····················· | ···················· | ···················· | ···················· |
| O | 5377 | hCoV-19/England/MILK-A7C07E/2020\|EPI_ISL_606340\|20 | ···················· | ···················· | ····················· | ···················· | ······T············· | ···················· |
| O | 5378 | hCoV-19/England/MILK-A7A82D/2020\|EPI_ISL_606341\|20 | ···················· | ···················· | ····················· | ···················· | ···················· | ···················· |
| O | 5379 | hCoV-19/England/MILK-A7C2A5/2020\|EPI_ISL_606342\|20 | ···················· | ···················· | ····················· | ···················· | ···················· | ···················· |
| O | 5380 | hCoV-19/England/MILK-A7AA36/2020\|EPI_ISL_606343\|20 | ···················· | ···················· | ····················· | ···················· | ···················· | ···················· |
| O | 5381 | hCoV-19/England/MILK-A7D63F/2020\|EPI_ISL_606344\|20 | ···················· | ···················· | ····················· | ···················· | ···················· | ···················· |
| O | 5382 | hCoV-19/England/MILK-A7DABB/2020\|EPI_ISL_606345\|20 | ···················· | ···················· | ····················· | ···················· | ···················· | ···················· |
| O | 5383 | hCoV-19/England/MILK-A7DC1F/2020\|EPI_ISL_606346\|20 | ···················· | ···················· | ····················· | ···················· | ···················· | ···················· |
| O | 5384 | hCoV-19/England/MILK-A7A6F6/2020\|EPI_ISL_606347\|20 | ···················· | ···················· | ····················· | ···················· | ···················· | ···················· |
| O | 5385 | hCoV-19/England/MILK-A7D12F/2020\|EPI_ISL_606348\|20 | ···················· | ···················· | ····················· | ···················· | ···················· | ···················· |
| O | 5386 | hCoV-19/England/MILK-A7C21E/2020\|EPI_ISL_606349\|20 | ···················· | ···················· | ····················· | ···················· | ···················· | ···················· |
| O | 5387 | hCoV-19/England/MILK-A7A80F/2020\|EPI_ISL_606350\|20 | ···················· | ···················· | ····················· | ···················· | ···················· | ···················· |
| O | 5388 | hCoV-19/England/MILK-A7C2E1/2020\|EPI_ISL_606351\|20 | ···················· | ···················· | ····················· | ···················· | ······T············· | ···················· |
| O | 5389 | hCoV-19/England/MILK-A7CADA/2020\|EPI_ISL_606352\|20 | ···················· | ···················· | ····················· | ···················· | ···················· | ···················· |
| O | 5390 | hCoV-19/England/MILK-A7CD4A/2020\|EPI_ISL_606354\|20 | ···················· | ···················· | ····················· | ···················· | ···················· | ···················· |
| O | 5391 | hCoV-19/England/MILK-A7C102/2020\|EPI_ISL_606355\|20 | ···················· | ···················· | ····················· | ···················· | ···················· | ···················· |
| O | 5392 | hCoV-19/England/MILK-A7DDB2/2020\|EPI_ISL_606356\|20 | ···················· | ···················· | ····················· | ···················· | ···················· | ···················· |
| O | 5393 | hCoV-19/England/MILK-A7C25A/2020\|EPI_ISL_606357\|20 | ···················· | ···················· | ····················· | ···················· | ···················· | ···················· |
| O | 5394 | hCoV-19/England/MILK-A7DD85/2020\|EPI_ISL_606358\|20 | ···················· | ···················· | ····················· | ···················· | ···················· | ···················· |
| O | 5395 | hCoV-19/England/MILK-A7C08D/2020\|EPI_ISL_606359\|20 | ···················· | ···················· | ····················· | ···················· | ···················· | ···················· |
| O | 5396 | hCoV-19/England/MILK-A7CA52/2020\|EPI_ISL_606361\|20 | ···················· | ···················· | ····················· | ···················· | ···················· | ···················· |
| O | 5397 | hCoV-19/England/MILK-A7A7C6/2020\|EPI_ISL_606362\|20 | ···················· | ···················· | ····················· | ···················· | ···················· | ···················· |
| O | 5398 | hCoV-19/England/MILK-A7D8A2/2020\|EPI_ISL_606363\|20 | ···················· | ···················· | ····················· | ···················· | ···················· | ···················· |
| O | 5399 | hCoV-19/England/MILK-A7DEEC/2020\|EPI_ISL_606364\|20 | ···················· | ···················· | ····················· | ···················· | ···················· | ···················· |
| O | 5400 | hCoV-19/England/MILK-A7D80C/2020\|EPI_ISL_606365\|20 | ···················· | ···················· | ····················· | ···················· | ···················· | ···················· |
| O | 5401 | hCoV-19/England/MILK-A7D5D8/2020\|EPI_ISL_606366\|20 | ···················· | ···················· | ····················· | ···················· | ···················· | ···················· |
| O | 5402 | hCoV-19/England/MILK-A7D550/2020\|EPI_ISL_606367\|20 | ···················· | ·A·················· | ····················· | ···················· | ···················· | ···················· |
| O | 5403 | hCoV-19/England/MILK-A7D1A7/2020\|EPI_ISL_606368\|20 | ···················· | ···················· | ····················· | ···················· | ···················· | ···················· |
| O | 5404 | hCoV-19/England/MILK-A7DC2E/2020\|EPI_ISL_606369\|20 | ···················· | ···················· | ····················· | ···················· | ···················· | ···················· |
| O | 5405 | hCoV-19/England/MILK-A7C418/2020\|EPI_ISL_606371\|20 | ···················· | ···················· | ····················· | ···················· | ···················· | ···················· |
| O | 5406 | hCoV-19/England/MILK-A7D7D2/2020\|EPI_ISL_606372\|20 | ···················· | ···················· | ····················· | ···················· | ···················· | ···················· |
| O | 5407 | hCoV-19/England/MILK-A7C2B4/2020\|EPI_ISL_606373\|20 | ···················· | ·A·················· | ····················· | ···················· | ···················· | ···················· |
| O | 5408 | hCoV-19/England/MILK-A7AE76/2020\|EPI_ISL_606376\|20 | ···················· | ···················· | ····················· | ···················· | ···················· | ···················· |
| O | 5409 | hCoV-19/England/MILK-A7D2B3/2020\|EPI_ISL_606377\|20 | ···················· | ···················· | ····················· | ···················· | ···················· | ···················· |
| O | 5410 | hCoV-19/England/MILK-A7D30B/2020\|EPI_ISL_606378\|20 | ···················· | ···················· | ····················· | ···················· | ···················· | ···················· |
| O | 5411 | hCoV-19/England/MILK-A7DAAC/2020\|EPI_ISL_606380\|20 | ···················· | ···················· | ····················· | ···················· | ···················· | ···················· |
| O | 5412 | hCoV-19/England/MILK-A7C32A/2020\|EPI_ISL_606382\|20 | ···················· | ···················· | ····················· | ···················· | ···················· | ···················· |
| O | 5413 | hCoV-19/England/MILK-A7D259/2020\|EPI_ISL_606383\|20 | ···················· | ···················· | ····················· | ···················· | ···················· | ···················· |
| O | 5414 | hCoV-19/England/MILK-A7D4CC/2020\|EPI_ISL_606384\|20 | ···················· | ···················· | ····················· | ···················· | ···················· | ···················· |
| O | 5415 | hCoV-19/England/MILK-A7D23B/2020\|EPI_ISL_606385\|20 | ···················· | ···················· | ····················· | ···················· | ···················· | ···················· |
| O | 5416 | hCoV-19/England/MILK-A7D778/2020\|EPI_ISL_606386\|20 | ···················· | ···················· | ····················· | ···················· | ······T············· | ···················· |
| O | 5417 | hCoV-19/England/MILK-A7CFEA/2020\|EPI_ISL_606387\|20 | ···················· | ···················· | ····················· | ···················· | ···················· | ···················· |
| O | 5418 | hCoV-19/England/MILK-A7D602/2020\|EPI_ISL_606388\|20 | ···················· | ···················· | ····················· | ···················· | ···················· | ···················· |
| O | 5419 | hCoV-19/England/MILK-A7C700/2020\|EPI_ISL_606389\|20 | ···················· | ···················· | ····················· | ···················· | ······T············· | ···················· |
| O | 5420 | hCoV-19/England/MILK-A7A76C/2020\|EPI_ISL_606390\|20 | ···················· | ···················· | ····················· | ···················· | ···················· | ···················· |
| O | 5421 | hCoV-19/England/MILK-A7D453/2020\|EPI_ISL_606391\|20 | ···················· | ···················· | ····················· | ···················· | ···················· | ···················· |
| O | 5422 | hCoV-19/England/MILK-A7D6C6/2020\|EPI_ISL_606392\|20 | ···················· | ···················· | ····················· | ···················· | ···················· | ···················· |
| O | 5423 | hCoV-19/England/MILK-A7D4F9/2020\|EPI_ISL_606393\|20 | ···················· | ···················· | ····················· | ···················· | ···················· | ···················· |
| O | 5424 | hCoV-19/England/MILK-A7C72E/2020\|EPI_ISL_606394\|20 | ···················· | ·A·················· | ····················· | ···················· | ···················· | ···A················ |
| O | 5425 | hCoV-19/England/MILK-A7C041/2020\|EPI_ISL_606395\|20 | ···················· | ···················· | ····················· | ···················· | ···················· | ···················· |
| O | 5426 | hCoV-19/England/MILK-A7C612/2020\|EPI_ISL_606396\|20 | ···················· | ···················· | ····················· | ···················· | ···················· | ···················· |
| O | 5427 | hCoV-19/England/MILK-A7D329/2020\|EPI_ISL_606397\|20 | ···················· | ···················· | ····················· | ···················· | ···················· | ···················· |
| O | 5428 | hCoV-19/England/MILK-A7DAE8/2020\|EPI_ISL_606398\|20 | ···················· | ···················· | ····················· | ···················· | ···················· | ···················· |
| O | 5429 | hCoV-19/England/MILK-A7D347/2020\|EPI_ISL_606399\|20 | ···················· | ···················· | ····················· | ···················· | ···················· | ···················· |
| O | 5430 | hCoV-19/England/MILK-A7DEDD/2020\|EPI_ISL_606400\|20 | ···················· | ···················· | ····················· | ···················· | ···················· | ···················· |
| O | 5431 | hCoV-19/England/MILK-A7DD94/2020\|EPI_ISL_606401\|20 | ···················· | ···················· | ····················· | ···················· | ···················· | ···················· |
| O | 5432 | hCoV-19/England/MILK-A7D3ED/2020\|EPI_ISL_606403\|20 | ···················· | ···················· | ····················· | ···················· | ···················· | ···················· |
| O | 5433 | hCoV-19/England/MILK-A7A78A/2020\|EPI_ISL_606405\|20 | ···················· | ···················· | ····················· | ···················· | ···················· | ···················· |
| O | 5434 | hCoV-19/England/MILK-A7C6F4/2020\|EPI_ISL_606406\|20 | ···················· | ···················· | ····················· | ···················· | ···················· | ···················· |

| | | | | | | | |
|---|---|---|---|---|---|---|---|
| O | 5435 hCoV-19/England/MILK-A7D7C3/2020\|EPI_ISL_606407\|20 | ···················· | ·A·················· | ···················· | ···················· | ···················· | ···················· |
| O | 5436 hCoV-19/England/MILK-A7CB7D/2020\|EPI_ISL_606409\|20 | ···················· | ···················· | ···················· | ···················· | ···················· | ···················· |
| O | 5437 hCoV-19/England/MILK-A7A702/2020\|EPI_ISL_606410\|20 | ···················· | ···················· | ···················· | ···················· | ······T············· | ···················· |
| O | 5438 hCoV-19/England/MILK-A7A68D/2020\|EPI_ISL_606411\|20 | ···················· | ···················· | ···················· | ···················· | ···················· | ···················· |
| O | 5439 hCoV-19/England/MILK-A7C0C9/2020\|EPI_ISL_606412\|20 | ···················· | ···················· | ···················· | ···················· | ······T············· | ···················· |
| O | 5440 hCoV-19/England/MILK-A7AD6A/2020\|EPI_ISL_606413\|20 | ···················· | ···················· | ···················· | ···················· | ···················· | ···················· |
| O | 5441 hCoV-19/England/MILK-A7AA90/2020\|EPI_ISL_606414\|20 | ···················· | ···················· | ···················· | ···················· | ···················· | ···················· |
| O | 5442 hCoV-19/England/MILK-A7A957/2020\|EPI_ISL_606417\|20 | ···················· | ···················· | ···················· | ···················· | ···················· | ···················· |
| O | 5443 hCoV-19/England/MILK-A7A975/2020\|EPI_ISL_606418\|20 | ···················· | ···················· | ···················· | ···················· | ···················· | ···················· |
| O | 5444 hCoV-19/England/MILK-A7C30C/2020\|EPI_ISL_606419\|20 | ···················· | ···················· | ···················· | ···················· | ······T············· | ···················· |
| O | 5445 hCoV-19/England/MILK-A7D374/2020\|EPI_ISL_606420\|20 | ···················· | ···················· | ···················· | ···················· | ···················· | ···················· |
| O | 5446 hCoV-19/England/MILK-A7AA45/2020\|EPI_ISL_606421\|20 | ···················· | ···················· | ···················· | ···················· | ···················· | ···················· |
| O | 5447 hCoV-19/England/MILK-A7D1E3/2020\|EPI_ISL_606423\|20 | ···················· | ···················· | ···················· | ···················· | ···················· | ···················· |
| O | 5448 hCoV-19/England/MILK-A7CA70/2020\|EPI_ISL_606426\|20 | ···················· | ···················· | ···················· | ···················· | ···················· | ···················· |
| O | 5449 hCoV-19/England/MILK-A7D541/2020\|EPI_ISL_606428\|20 | ···················· | ···················· | ···················· | ···················· | ···················· | ···················· |
| O | 5450 hCoV-19/England/MILK-A7DC6A/2020\|EPI_ISL_606429\|20 | ···················· | ···················· | ···················· | ···················· | ···················· | ···················· |
| O | 5451 hCoV-19/England/MILK-A7D14D/2020\|EPI_ISL_606430\|20 | ···················· | ···················· | ···················· | ···················· | ···················· | ···················· |
| O | 5452 hCoV-19/England/MILK-A7D6E4/2020\|EPI_ISL_606431\|20 | ···················· | ···················· | ···················· | ···················· | ······T············· | ···················· |
| O | 5453 hCoV-19/England/MILK-A7ADA6/2020\|EPI_ISL_606432\|20 | ···················· | ···················· | ···················· | ···················· | ···················· | ···················· |
| O | 5454 hCoV-19/England/MILK-A7D022/2020\|EPI_ISL_606433\|20 | ···················· | ···················· | ···················· | ···················· | ···················· | ···················· |
| O | 5455 hCoV-19/England/MILK-A7A90C/2020\|EPI_ISL_606434\|20 | ···················· | ···················· | ···················· | ···················· | ···················· | ···················· |
| O | 5456 hCoV-19/England/MILK-A7AAEB/2020\|EPI_ISL_606435\|20 | ···················· | ···················· | ···················· | ···················· | ···················· | ···················· |
| O | 5457 hCoV-19/England/MILK-A7D417/2020\|EPI_ISL_606436\|20 | ···················· | ···················· | ···················· | ···················· | ···················· | ···················· |
| O | 5458 hCoV-19/England/MILK-A7C1E4/2020\|EPI_ISL_606437\|20 | ···················· | ·A·················· | ···················· | ···················· | ···················· | ··A················· |
| O | 5459 hCoV-19/England/MILK-A7C296/2020\|EPI_ISL_606438\|20 | ···················· | ···················· | ···················· | ···················· | ···················· | ···················· |
| O | 5460 hCoV-19/England/MILK-A7C111/2020\|EPI_ISL_606439\|20 | ···················· | ·A·················· | ···················· | ···················· | ···················· | ··A················· |
| O | 5461 hCoV-19/England/MILK-A7D198/2020\|EPI_ISL_606440\|20 | ···················· | ···················· | ···················· | ···················· | ···················· | ···················· |
| O | 5462 hCoV-19/England/MILK-A7C3A2/2020\|EPI_ISL_606441\|20 | ···················· | ···················· | ···················· | ···················· | ···················· | ···················· |
| O | 5463 hCoV-19/England/MILK-A7D3FC/2020\|EPI_ISL_606442\|20 | ···················· | ···················· | ···················· | ···················· | ···················· | ···················· |
| O | 5464 hCoV-19/England/MILK-A7AF46/2020\|EPI_ISL_606443\|20 | ···················· | ···················· | ···················· | ···················· | ···················· | ···················· |
| O | 5465 hCoV-19/England/MILK-A7C65E/2020\|EPI_ISL_606444\|20 | ···················· | ···················· | ···················· | ···················· | ···················· | ···················· |
| O | 5466 hCoV-19/England/MILK-A7AADC/2020\|EPI_ISL_606445\|20 | ···················· | ···················· | ···················· | ···················· | ···················· | ···················· |
| O | 5467 hCoV-19/England/MILK-A7D24A/2020\|EPI_ISL_606446\|20 | ···················· | ···················· | ···················· | ···················· | ···················· | ···················· |
| O | 5468 hCoV-19/England/MILK-A7A799/2020\|EPI_ISL_606447\|20 | ···················· | ···················· | ···················· | ···················· | ···················· | ···················· |
| O | 5469 hCoV-19/England/MILK-A7D2A4/2020\|EPI_ISL_606448\|20 | ···················· | ···················· | ···················· | ···················· | ···················· | ···················· |
| O | 5470 hCoV-19/England/MILK-A7C3DF/2020\|EPI_ISL_606449\|20 | ···················· | ···················· | ···················· | ···················· | ···················· | ···················· |
| O | 5471 hCoV-19/England/MILK-A7D523/2020\|EPI_ISL_606450\|20 | ···················· | ·A·················· | ···················· | ···················· | ···················· | ··A················· |
| O | 5472 hCoV-19/England/MILK-A7C09C/2020\|EPI_ISL_606451\|20 | ···················· | ···················· | ···················· | ···················· | ···················· | ···················· |
| O | 5473 hCoV-19/England/MILK-A7D70F/2020\|EPI_ISL_606453\|20 | ···················· | ···················· | ···················· | ···················· | ···················· | ···················· |
| O | 5474 hCoV-19/England/MILK-A7C1F3/2020\|EPI_ISL_606454\|20 | ···················· | ···················· | ···················· | ···················· | ···················· | ···················· |
| O | 5475 hCoV-19/England/MILK-A7DECE/2020\|EPI_ISL_606455\|20 | ···················· | ···················· | ···················· | ···················· | ···················· | ···················· |
| O | 5476 hCoV-19/England/MILK-A7D857/2020\|EPI_ISL_606456\|20 | ···················· | ···················· | ···················· | ···················· | ···················· | ···················· |
| O | 5477 hCoV-19/England/MILK-A7AE49/2020\|EPI_ISL_606457\|20 | ···················· | ···················· | ···················· | ···················· | ···················· | ···················· |
| O | 5478 hCoV-19/England/MILK-A7DC3D/2020\|EPI_ISL_606458\|20 | ···················· | ···················· | ···················· | ···················· | ···················· | ···················· |
| O | 5479 hCoV-19/England/ALDP-A6EA26/2020\|EPI_ISL_607670\|20 | ···················· | ···················· | ···················· | ···················· | ···················· | ···················· |
| O | 5480 hCoV-19/England/ALDP-A704BA/2020\|EPI_ISL_607671\|20 | ···················· | ···················· | ···················· | ···················· | ···················· | ···················· |
| O | 5481 hCoV-19/England/ALDP-A6DEC1/2020\|EPI_ISL_607672\|20 | ···················· | ···················· | ···················· | ···················· | ···················· | ···················· |
| O | 5482 hCoV-19/England/ALDP-A6F5F7/2020\|EPI_ISL_607673\|20 | ···················· | ···················· | ···················· | ···················· | ···················· | ···················· |
| O | 5483 hCoV-19/England/ALDP-A706A5/2020\|EPI_ISL_607674\|20 | ···················· | ···················· | ···················· | ···················· | ···················· | ···················· |
| O | 5484 hCoV-19/England/ALDP-A6DEFE/2020\|EPI_ISL_607676\|20 | ···················· | ···················· | ···················· | ···················· | ···················· | ···················· |
| O | 5485 hCoV-19/England/ALDP-A70520/2020\|EPI_ISL_607677\|20 | ···················· | ···················· | ···················· | ···················· | ···················· | ···················· |
| O | 5486 hCoV-19/England/ALDP-A6E1D6/2020\|EPI_ISL_607678\|20 | ···················· | ···················· | ···················· | ···················· | ···················· | ···················· |
| O | 5487 hCoV-19/England/ALDP-A6EFEB/2020\|EPI_ISL_607679\|20 | ···················· | ···················· | ···················· | ···················· | ···················· | ···················· |
| O | 5488 hCoV-19/England/ALDP-A6F111/2020\|EPI_ISL_607681\|20 | ···················· | ···················· | ···················· | ···················· | ···················· | ···················· |
| O | 5489 hCoV-19/England/ALDP-A6F69A/2020\|EPI_ISL_607682\|20 | ···················· | ···················· | ···················· | ···················· | ···················· | ···················· |
| O | 5490 hCoV-19/England/ALDP-A6EACC/2020\|EPI_ISL_607683\|20 | ···················· | ···················· | ···················· | ···················· | ···················· | ···················· |
| O | 5491 hCoV-19/England/ALDP-A7065A/2020\|EPI_ISL_607684\|20 | ···················· | ···················· | ···················· | ···················· | ···················· | ···················· |
| O | 5492 hCoV-19/England/ALDP-A702B0/2020\|EPI_ISL_607685\|20 | ···················· | ···················· | ···················· | ···················· | ···················· | ···················· |
| O | 5493 hCoV-19/England/ALDP-A6F472/2020\|EPI_ISL_607686\|20 | ···················· | ···················· | ···················· | ···················· | ···················· | ···················· |
| O | 5494 hCoV-19/England/ALDP-A6D8C3/2020\|EPI_ISL_607687\|20 | ···················· | ···················· | ···················· | ···················· | ···················· | ···················· |
| O | 5495 hCoV-19/England/ALDP-A6DB7F/2020\|EPI_ISL_607688\|20 | ···················· | ···················· | ···················· | ···················· | ···················· | ···················· |
| O | 5496 hCoV-19/England/ALDP-A6E929/2020\|EPI_ISL_607689\|20 | ···················· | ·A·················· | ···················· | ···················· | ···················· | ··A················· |
| O | 5497 hCoV-19/England/ALDP-A6E428/2020\|EPI_ISL_607691\|20 | ···················· | ···················· | ···················· | ···················· | ···················· | ···················· |
| O | 5498 hCoV-19/England/ALDP-A6F06F/2020\|EPI_ISL_607692\|20 | ···················· | ···················· | ···················· | ···················· | ···················· | ···················· |
| O | 5499 hCoV-19/England/ALDP-A6E0AC/2020\|EPI_ISL_607693\|20 | ···················· | ···················· | ···················· | ···················· | ···················· | ···················· |
| O | 5500 hCoV-19/England/ALDP-A6ECE4/2020\|EPI_ISL_607694\|20 | ···················· | ···················· | ···················· | ···················· | ···················· | ···················· |
| O | 5501 hCoV-19/England/ALDP-A6F09C/2020\|EPI_ISL_607695\|20 | ···················· | ···················· | ···················· | ···················· | ···················· | ···················· |
| O | 5502 hCoV-19/England/ALDP-A6E033/2020\|EPI_ISL_607697\|20 | ···················· | ···················· | ···················· | ···················· | ···················· | ···················· |
| O | 5503 hCoV-19/England/ALDP-A6F436/2020\|EPI_ISL_607699\|20 | ···················· | ···················· | ···················· | ···················· | ······T············· | ···················· |
| O | 5504 hCoV-19/England/ALDP-A7070C/2020\|EPI_ISL_607708\|20 | ···················· | ···················· | ···················· | ···················· | ···················· | ···················· |
| O | 5505 hCoV-19/England/ALDP-A6DF46/2020\|EPI_ISL_607709\|20 | ···················· | ···················· | ···················· | ···················· | ···················· | ···················· |
| O | 5506 hCoV-19/England/ALDP-A704D8/2020\|EPI_ISL_607711\|20 | ···················· | ···················· | ···················· | ···················· | ···················· | ···················· |
| O | 5507 hCoV-19/England/ALDP-A70441/2020\|EPI_ISL_607712\|20 | ···················· | ···················· | ···················· | ···················· | ···················· | ···················· |
| O | 5508 hCoV-19/England/ALDP-A6E72F/2020\|EPI_ISL_607714\|20 | ···················· | ···················· | ···················· | ···················· | ···················· | ···················· |
| O | 5509 hCoV-19/England/ALDP-A6F102/2020\|EPI_ISL_607715\|20 | ···················· | ···················· | ···················· | ···················· | ···················· | ···················· |
| O | 5510 hCoV-19/England/ALDP-A6F07E/2020\|EPI_ISL_607718\|20 | ···················· | ···················· | ···················· | ···················· | ···················· | ···················· |
| O | 5511 hCoV-19/England/ALDP-A6F120/2020\|EPI_ISL_607719\|20 | ···················· | ···················· | ···················· | ···················· | ···················· | ···················· |
| O | 5512 hCoV-19/England/ALDP-A6E7B6/2020\|EPI_ISL_607720\|20 | ···················· | ···················· | ···················· | ···················· | ···················· | ···················· |
| O | 5513 hCoV-19/England/ALDP-A6D896/2020\|EPI_ISL_607721\|20 | ···················· | ···················· | ···················· | ···················· | ···················· | ···················· |
| O | 5514 hCoV-19/England/ALDP-A706C3/2020\|EPI_ISL_607722\|20 | ···················· | ···················· | ···················· | ···················· | ···················· | ···················· |

| | | | | | | | |
|---|---|---|---|---|---|---|---|
| O | 5515 hCoV-19/England/ALDP-A6E0BB/2020\|EPI_ISL_607723\|20 | ···················· | ···················· | ···················· | ···················· | ···················· | ···················· |
| O | 5516 hCoV-19/England/ALDP-A6EABD/2020\|EPI_ISL_607724\|20 | ···················· | ···················· | ···················· | ···················· | ···················· | ···················· |
| O | 5517 hCoV-19/England/ALDP-A6F1A8/2020\|EPI_ISL_607726\|20 | ···················· | ···················· | ···················· | ···················· | ···················· | ···················· |
| O | 5518 hCoV-19/England/ALDP-A7063C/2020\|EPI_ISL_607727\|20 | ···················· | ···················· | ···················· | ···················· | ···················· | ···················· |
| O | 5519 hCoV-19/England/ALDP-A6F023/2020\|EPI_ISL_607728\|20 | ···················· | ···················· | ···················· | ···················· | ···················· | ···················· |
| O | 5520 hCoV-19/England/ALDP-A6DF0A/2020\|EPI_ISL_607729\|20 | ···················· | ···················· | ···················· | ···················· | ···················· | ···················· |
| O | 5521 hCoV-19/England/ALDP-A6F041/2020\|EPI_ISL_607730\|20 | ···················· | ···················· | ···················· | ···················· | ···················· | ···················· |
| O | 5522 hCoV-19/England/ALDP-A6E9B0/2020\|EPI_ISL_607731\|20 | ···················· | ···················· | ···················· | ···················· | ···················· | ···················· |
| O | 5523 hCoV-19/England/ALDP-A6E886/2020\|EPI_ISL_607732\|20 | ···················· | ·A·················· | ···················· | ···················· | ···················· | ··A················· |
| O | 5524 hCoV-19/England/ALDP-A6F66D/2020\|EPI_ISL_607733\|20 | ···················· | ···················· | ···················· | ···················· | ···················· | ···················· |
| O | 5525 hCoV-19/England/ALDP-A6E74D/2020\|EPI_ISL_607734\|20 | ···················· | ···················· | ···················· | ···················· | ···················· | ···················· |
| O | 5526 hCoV-19/England/ALDP-A6E446/2020\|EPI_ISL_607735\|20 | ···················· | ···················· | ···················· | ···················· | ···················· | ···················· |
| O | 5527 hCoV-19/England/ALDP-A7039F/2020\|EPI_ISL_607736\|20 | ···················· | ···················· | ···················· | ···················· | ···················· | ···················· |
| O | 5528 hCoV-19/England/ALDP-A6F4EB/2020\|EPI_ISL_607737\|20 | ···················· | ···················· | ···················· | ···················· | ···················· | ···················· |
| O | 5529 hCoV-19/England/ALDP-A6F0E7/2020\|EPI_ISL_607738\|20 | ···················· | ···················· | ···················· | ···················· | ···················· | ···················· |
| O | 5530 hCoV-19/England/ALDP-A6DEA3/2020\|EPI_ISL_607739\|20 | ···················· | ···················· | ···················· | ···················· | ···················· | ···················· |
| O | 5531 hCoV-19/England/ALDP-A70317/2020\|EPI_ISL_607740\|20 | ···················· | ···················· | ···················· | ···················· | ···················· | ···················· |
| O | 5532 hCoV-19/England/ALDP-A6F506/2020\|EPI_ISL_607741\|20 | ···················· | ···················· | ···················· | ···················· | ···················· | ···················· |
| O | 5533 hCoV-19/England/ALDP-A6F991/2020\|EPI_ISL_607742\|20 | ···················· | ···················· | ···················· | ···················· | ···················· | ···················· |
| O | 5534 hCoV-19/England/ALDP-A6E877/2020\|EPI_ISL_607743\|20 | ···················· | ···················· | ···················· | ···················· | ···················· | ···················· |
| O | 5535 hCoV-19/England/ALDP-A704F6/2020\|EPI_ISL_607744\|20 | ···················· | ·A·················· | ···················· | ···················· | ···················· | ···················· |
| O | 5536 hCoV-19/England/ALDP-A6F982/2020\|EPI_ISL_607745\|20 | ···················· | ···················· | ···················· | ···················· | ···················· | ···················· |
| O | 5537 hCoV-19/England/ALDP-A6FA52/2020\|EPI_ISL_607746\|20 | ···················· | ···················· | ···················· | ···················· | ···················· | ···················· |
| O | 5538 hCoV-19/England/ALDP-A6DB60/2020\|EPI_ISL_607747\|20 | ···················· | ···················· | ···················· | ···················· | ···················· | ···················· |
| O | 5539 hCoV-19/England/ALDP-A6DA27/2020\|EPI_ISL_607748\|20 | ···················· | ···················· | ···················· | ···················· | ···················· | ···················· |
| O | 5540 hCoV-19/England/ALDP-A6E965/2020\|EPI_ISL_607749\|20 | ···················· | ···················· | ···················· | ···················· | ···················· | ···················· |
| O | 5541 hCoV-19/England/ALDP-A704E7/2020\|EPI_ISL_607750\|20 | ···················· | ···················· | ···················· | ···················· | ···················· | ···················· |
| O | 5542 hCoV-19/England/ALDP-A6E491/2020\|EPI_ISL_607751\|20 | ···················· | ···················· | ···················· | ···················· | ···················· | ···················· |
| O | 5543 hCoV-19/England/ALDP-A6F25A/2020\|EPI_ISL_607752\|20 | ···················· | ···················· | ···················· | ···················· | ···················· | ···················· |
| O | 5544 hCoV-19/England/ALDP-A703CC/2020\|EPI_ISL_607753\|20 | ···················· | ···················· | ···················· | ···················· | ···················· | ···················· |
| O | 5545 hCoV-19/England/ALDP-A6E895/2020\|EPI_ISL_607754\|20 | ···················· | ···················· | ···················· | ···················· | ···················· | ···················· |
| O | 5546 hCoV-19/England/ALDP-A6DC12/2020\|EPI_ISL_607755\|20 | ···················· | ···················· | ···················· | ···················· | ···················· | ···················· |
| O | 5547 hCoV-19/England/ALDP-A705F3/2020\|EPI_ISL_607756\|20 | ···················· | ···················· | ···················· | ···················· | ···················· | ···················· |
| O | 5548 hCoV-19/England/ALDP-A6DFCE/2020\|EPI_ISL_607757\|20 | ···················· | ···················· | ···················· | ···················· | ···················· | ···················· |
| O | 5549 hCoV-19/England/ALDP-A6F0D8/2020\|EPI_ISL_607758\|20 | ···················· | ···················· | ···················· | ···················· | ···················· | ···················· |
| O | 5550 hCoV-19/England/ALDP-A6E75C/2020\|EPI_ISL_607759\|20 | ···················· | ···················· | ···················· | ···················· | ···················· | ···················· |
| O | 5551 hCoV-19/England/ALDP-A6DABE/2020\|EPI_ISL_607760\|20 | ···················· | ···················· | ···················· | ···················· | ···················· | ···················· |
| O | 5552 hCoV-19/England/ALDP-A6F9EC/2020\|EPI_ISL_607761\|20 | ···················· | ···················· | ···················· | ···················· | ···················· | ···················· |
| O | 5553 hCoV-19/England/ALDP-A70687/2020\|EPI_ISL_607762\|20 | ···················· | ···················· | ···················· | ···················· | ···················· | ···················· |
| O | 5554 hCoV-19/England/ALDP-A70380/2020\|EPI_ISL_607763\|20 | ···················· | ···················· | ···················· | ···················· | ···················· | ···················· |
| O | 5555 hCoV-19/England/ALDP-A6E956/2020\|EPI_ISL_607764\|20 | ···················· | ···················· | ···················· | ···················· | ···················· | ···················· |
| O | 5556 hCoV-19/England/ALDP-A6DC30/2020\|EPI_ISL_607766\|20 | ···················· | ·A·················· | ···················· | ···················· | ···················· | ···················· |
| O | 5557 hCoV-19/England/ALDP-A6E40A/2020\|EPI_ISL_607768\|20 | ···················· | ···················· | ···················· | ···················· | ···················· | ···················· |
| O | 5558 hCoV-19/England/ALDP-A6F621/2020\|EPI_ISL_607769\|20 | ···················· | ···················· | ···················· | ···················· | ···················· | ···················· |
| O | 5559 hCoV-19/England/ALDP-A6E042/2020\|EPI_ISL_607770\|20 | ···················· | ···················· | ···················· | ···················· | ···················· | ···················· |
| O | 5560 hCoV-19/England/ALDP-A6F65E/2020\|EPI_ISL_607771\|20 | ···················· | ···················· | ···················· | ···················· | ···················· | ···················· |
| O | 5561 hCoV-19/England/ALDP-A6DE76/2020\|EPI_ISL_607772\|20 | ···················· | ···················· | ···················· | ···················· | ···················· | ···················· |
| O | 5562 hCoV-19/England/ALDP-A6F4AF/2020\|EPI_ISL_607774\|20 | ···················· | ···················· | ···················· | ···················· | ···················· | ···················· |
| O | 5563 hCoV-19/England/ALDP-A6ED3C/2020\|EPI_ISL_607776\|20 | ····W··············· | ···················· | ···················· | ···················· | ···················· | ···················· |
| O | 5564 hCoV-19/England/ALDP-A70344/2020\|EPI_ISL_607777\|20 | ···················· | ···················· | ···················· | ···················· | ···················· | ···················· |
| O | 5565 hCoV-19/England/ALDP-A6DF28/2020\|EPI_ISL_607778\|20 | ···················· | ···················· | ···················· | ···················· | ···················· | ···················· |
| O | 5566 hCoV-19/England/ALDP-A7049C/2020\|EPI_ISL_607779\|20 | ···················· | ···················· | ···················· | ···················· | ···················· | ···················· |
| O | 5567 hCoV-19/England/ALDP-A6E8FF/2020\|EPI_ISL_607781\|20 | ···················· | ···················· | ···················· | ···················· | ···················· | ···················· |
| O | 5568 hCoV-19/England/ALDP-A6F463/2020\|EPI_ISL_607782\|20 | ···················· | ···················· | ···················· | ···················· | ···················· | ···················· |
| O | 5569 hCoV-19/England/ALDP-A6DFDD/2020\|EPI_ISL_607783\|20 | ···················· | ···················· | ···················· | ···················· | ···················· | ···················· |
| O | 5570 hCoV-19/England/ALDP-A6ED87/2020\|EPI_ISL_607784\|20 | ···················· | ···················· | ···················· | ···················· | ···················· | ···················· |
| O | 5571 hCoV-19/England/ALDP-A6E015/2020\|EPI_ISL_607785\|20 | ···················· | ·A·················· | ···················· | ···················· | ···················· | ···················· |
| O | 5572 hCoV-19/England/ALDP-A702CF/2020\|EPI_ISL_607786\|20 | ···················· | ···················· | ···················· | ···················· | ···················· | ···················· |
| O | 5573 hCoV-19/England/ALDP-A6F630/2020\|EPI_ISL_607787\|20 | ···················· | ···················· | ···················· | ···················· | ···················· | ···················· |
| O | 5574 hCoV-19/England/ALDP-A6FA43/2020\|EPI_ISL_607788\|20 | ···················· | ···················· | ···················· | ···················· | ···················· | ···················· |
| O | 5575 hCoV-19/England/ALDP-A6EF90/2020\|EPI_ISL_607789\|20 | ···················· | ···················· | ···················· | ···················· | ···················· | ···················· |
| O | 5578 hCoV-19/England/BIRM-65DD5/2020\|EPI_ISL_612229\|202 | ···················· | ···················· | ···················· | ···················· | ···················· | ···················· |
| O | 5579 hCoV-19/England/BIRM-65DF3/2020\|EPI_ISL_612230\|202 | ···················· | ···················· | ···················· | ···················· | ···················· | ···················· |
| O | 5580 hCoV-19/England/BIRM-65E2D/2020\|EPI_ISL_612231\|202 | ···················· | ···················· | ···················· | ···················· | ···················· | ···················· |
| O | 5581 hCoV-19/England/BIRM-65E3C/2020\|EPI_ISL_612232\|202 | ···················· | ···················· | ···················· | ···················· | ···················· | ···················· |
| O | 5582 hCoV-19/England/BIRM-65E4B/2020\|EPI_ISL_612233\|202 | ···················· | ···················· | ···················· | ···················· | ···················· | ···················· |
| O | 5583 hCoV-19/England/BIRM-65F1B/2020\|EPI_ISL_612234\|202 | ···················· | ···················· | ···················· | ···················· | ···················· | ···················· |
| O | 5585 hCoV-19/England/BIRM-66114/2020\|EPI_ISL_612236\|202 | ···················· | ···················· | ···················· | ···················· | ···················· | ···················· |
| O | 5586 hCoV-19/England/BIRM-66123/2020\|EPI_ISL_612237\|202 | ···················· | ···················· | ···················· | ···················· | ···················· | ···················· |
| O | 5587 hCoV-19/England/BIRM-66132/2020\|EPI_ISL_612238\|202 | ···················· | ···················· | ···················· | ···················· | ···················· | ···················· |
| O | 5588 hCoV-19/England/BIRM-6616F/2020\|EPI_ISL_612239\|202 | ···················· | ···················· | ···················· | ···················· | ···················· | ···················· |
| O | 5589 hCoV-19/England/BIRM-6617E/2020\|EPI_ISL_612240\|202 | ···················· | ···················· | ···················· | ···················· | ···················· | ···················· |
| O | 5590 hCoV-19/England/BIRM-6618D/2020\|EPI_ISL_612241\|202 | ···················· | ···················· | ···················· | ···················· | ···················· | ···················· |
| O | 5591 hCoV-19/England/BIRM-6619C/2020\|EPI_ISL_612242\|202 | ···················· | ·A·················· | ···················· | ···················· | ···················· | ··A················· |
| O | 5592 hCoV-19/England/BIRM-661AB/2020\|EPI_ISL_612243\|202 | ···················· | ···················· | ···················· | ···················· | ···················· | ···················· |
| O | 5593 hCoV-19/England/BIRM-661BA/2020\|EPI_ISL_612244\|202 | ···················· | ···················· | ···················· | ···················· | ···················· | ···················· |
| O | 5594 hCoV-19/England/BIRM-661D8/2020\|EPI_ISL_612245\|202 | ···················· | ···················· | ···················· | ···················· | ···················· | ···················· |
| O | 5595 hCoV-19/England/BIRM-661F6/2020\|EPI_ISL_612246\|202 | ···················· | ···················· | ···················· | ···················· | ···················· | ···················· |
| O | 5596 hCoV-19/England/BIRM-66202/2020\|EPI_ISL_612247\|202 | ···················· | ···················· | ···················· | ···················· | ···················· | ···················· |
| O | 5597 hCoV-19/England/BIRM-66211/2020\|EPI_ISL_612248\|202 | ···················· | ···················· | ···················· | ···················· | ···················· | ···················· |

| | | | | | | | |
|---|---|---|---|---|---|---|---|
| O | 5598 hCoV-19/England/BIRM-66220/2020\|EPI_ISL_612249\|202 | .................... | .................... | .................... | .................... | .................... | .................... |
| O | 5599 hCoV-19/England/BIRM-6625D/2020\|EPI_ISL_612250\|202 | .................... | .................... | .................... | .................... | .................... | .................... |
| O | 5600 hCoV-19/England/BIRM-6627B/2020\|EPI_ISL_612251\|202 | .................... | .................... | .................... | .................... | .................... | .................... |
| O | 5601 hCoV-19/England/BIRM-662A8/2020\|EPI_ISL_612252\|202 | .................... | .................... | .................... | .................... | .................... | .................... |
| O | 5602 hCoV-19/England/BIRM-662B7/2020\|EPI_ISL_612253\|202 | .................... | .................... | .................... | .................... | .................... | .................... |
| O | 5603 hCoV-19/England/BIRM-662D5/2020\|EPI_ISL_612254\|202 | .................... | .................... | .................... | .................... | .................... | .................... |
| O | 5604 hCoV-19/England/BIRM-662E4/2020\|EPI_ISL_612255\|202 | .................... | .................... | .................... | .................... | .................... | .................... |
| O | 5605 hCoV-19/England/BIRM-662F3/2020\|EPI_ISL_612256\|202 | .................... | .................... | .................... | .................... | .................... | .................... |
| O | 5606 hCoV-19/England/BIRM-6631E/2020\|EPI_ISL_612257\|202 | .................... | .................... | .................... | .................... | .................... | .................... |
| O | 5607 hCoV-19/England/BIRM-6633C/2020\|EPI_ISL_612258\|202 | .................... | .................... | .................... | .................... | .................... | .................... |
| O | 5609 hCoV-19/France/BRE-BR9060/2020\|EPI_ISL_613549\|2020 | .................... | .................... | .................... | .................... | .................... | .................... |
| O | 5610 hCoV-19/Italy/SAR-ASSL-01/2020\|EPI_ISL_613560\|2020 | .................... | .................... | .................... | .................... | .................... | .................... |
| O | 5611 hCoV-19/Brazil/SP-Pat14/2020\|EPI_ISL_613563\|2020-0 | .................... | .................... | .................... | .................... | .................... | .................... |
| O | 5612 hCoV-19/Brazil/SP-Pat22/2020\|EPI_ISL_613564\|2020-0 | .................... | .................... | .................... | .................... | .................... | .................... |
| O | 5613 hCoV-19/Gibraltar/204421247/2020\|EPI_ISL_613570\|20 | .................... | .A.................. | .................... | .................... | .................... | ...A................ |
| O | 5614 hCoV-19/England/204260069/2020\|EPI_ISL_613571\|2020 | .................... | .................... | .................... | .................... | .................... | .................... |
| O | 5615 hCoV-19/England/204260070/2020\|EPI_ISL_613572\|2020 | .................... | .................... | .................... | .................... | .................... | .................... |
| O | 5616 hCoV-19/England/204260089/2020\|EPI_ISL_613573\|2020 | .................... | .................... | .................... | .................... | .................... | .................... |
| O | 5617 hCoV-19/England/204260091/2020\|EPI_ISL_613574\|2020 | .................... | .................... | .................... | .................... | .................... | .................... |
| O | 5618 hCoV-19/England/204260092/2020\|EPI_ISL_613575\|2020 | .................... | .................... | .................... | .................... | .................... | .................... |
| O | 5619 hCoV-19/England/204260923/2020\|EPI_ISL_613576\|2020 | .................... | .................... | .................... | .................... | .................... | .................... |
| O | 5620 hCoV-19/England/204261202/2020\|EPI_ISL_613577\|2020 | .................... | .................... | .................... | .................... | .................... | .................... |
| O | 5621 hCoV-19/England/204261268/2020\|EPI_ISL_613578\|2020 | .................... | .................... | .................... | .................... | .................... | .................... |
| O | 5622 hCoV-19/England/204261269/2020\|EPI_ISL_613579\|2020 | .................... | .................... | .................... | .................... | .................... | .................... |
| O | 5623 hCoV-19/England/204261273/2020\|EPI_ISL_613580\|2020 | .................... | .................... | .................... | .................... | .................... | .................... |
| O | 5624 hCoV-19/England/204261275/2020\|EPI_ISL_613581\|2020 | .................... | .................... | .................... | .................... | .................... | .................... |
| O | 5625 hCoV-19/England/204261293/2020\|EPI_ISL_613582\|2020 | .................... | .................... | .................... | .................... | .................... | .................... |
| O | 5626 hCoV-19/England/204261294/2020\|EPI_ISL_613583\|2020 | .................... | .................... | .................... | .................... | .................... | .................... |
| O | 5627 hCoV-19/England/204261295/2020\|EPI_ISL_613584\|2020 | .................... | .................... | .................... | .................... | .................... | .................... |
| O | 5628 hCoV-19/England/204261303/2020\|EPI_ISL_613585\|2020 | .................... | .................... | .................... | .................... | .................... | .................... |
| O | 5629 hCoV-19/England/204261306/2020\|EPI_ISL_613586\|2020 | .................... | .................... | .................... | .................... | .................... | .................... |
| O | 5630 hCoV-19/England/204261316/2020\|EPI_ISL_613587\|2020 | .................... | .................... | .................... | .................... | .................... | .................... |
| O | 5631 hCoV-19/England/204261330/2020\|EPI_ISL_613588\|2020 | .................... | .................... | .................... | .................... | .................... | .................... |
| O | 5632 hCoV-19/England/204261331/2020\|EPI_ISL_613589\|2020 | .................... | .................... | .................... | .................... | .................... | .................... |
| O | 5633 hCoV-19/England/MILK-A7AA54/2020\|EPI_ISL_606301\|20 | .................... | .................... | .................... | .................... | .................... | .................... |
| O | 5634 hCoV-19/England/MILK-A7DD3A/2020\|EPI_ISL_606302\|20 | .................... | .................... | .................... | .................... | .................... | .................... |
| O | 5635 hCoV-19/England/MILK-A7C0D8/2020\|EPI_ISL_606303\|20 | .................... | .................... | .................... | .................... | .................... | .................... |
| O | 5636 hCoV-19/England/MILK-A7D2E0/2020\|EPI_ISL_606304\|20 | .................... | .A.................. | .................... | .................... | .................... | ...A................ |
| O | 5637 hCoV-19/England/MILK-A7DA9D/2020\|EPI_ISL_606305\|20 | .................... | .................... | .................... | .................... | .................... | .................... |
| O | 5638 hCoV-19/England/MILK-A7D7A5/2020\|EPI_ISL_606306\|20 | .................... | .................... | .................... | .................... | .................... | .................... |
| O | 5639 hCoV-19/England/MILK-A7D3A1/2020\|EPI_ISL_606307\|20 | .................... | ......A............. | .................... | .................... | .................... | .................... |
| O | 5640 hCoV-19/England/MILK-A7C490/2020\|EPI_ISL_606309\|20 | .................... | .................... | .................... | .................... | .................... | .................... |
| O | 5641 hCoV-19/England/MILK-A7C5F7/2020\|EPI_ISL_606310\|20 | .................... | .................... | .................... | .................... | .................... | .................... |
| O | 5642 hCoV-19/England/MILK-A7AB15/2020\|EPI_ISL_606312\|20 | .................... | .................... | .................... | .................... | .................... | .................... |
| O | 5643 hCoV-19/England/MILK-A7D59C/2020\|EPI_ISL_606313\|20 | .................... | .................... | .................... | .................... | .................... | .................... |
| O | 5644 hCoV-19/England/MILK-A7A7B7/2020\|EPI_ISL_606315\|20 | .................... | .................... | .................... | .................... | .................... | .................... |
| O | 5645 hCoV-19/England/MILK-A7C2C3/2020\|EPI_ISL_606317\|20 | .................... | .................... | .................... | .................... | .................... | .................... |
| O | 5646 hCoV-19/England/MILK-A7D295/2020\|EPI_ISL_606318\|20 | .................... | .................... | .................... | .................... | ......T............. | .................... |
| O | 5647 hCoV-19/England/MILK-A7C1D5/2020\|EPI_ISL_606322\|20 | .................... | .................... | .................... | .................... | .................... | .................... |
| O | 5648 hCoV-19/England/MILK-A7D6F3/2020\|EPI_ISL_606323\|20 | .................... | .................... | .................... | .................... | .................... | .................... |
| O | 5649 hCoV-19/England/MILK-A7D383/2020\|EPI_ISL_606324\|20 | .................... | .................... | .................... | .................... | .................... | .................... |
| O | 5650 hCoV-19/England/MILK-A7C287/2020\|EPI_ISL_606325\|20 | .................... | .................... | .................... | .................... | .................... | .................... |
| O | 5651 hCoV-19/England/MILK-A7DAD9/2020\|EPI_ISL_606326\|20 | .................... | .................... | .................... | .................... | .................... | .................... |
| O | 5652 hCoV-19/England/MILK-A7C050/2020\|EPI_ISL_606327\|20 | .................... | .................... | .................... | .................... | .................... | .................... |
| O | 5653 hCoV-19/England/MILK-A7D74B/2020\|EPI_ISL_606328\|20 | .................... | .................... | .................... | .................... | .................... | .................... |
| O | 5654 hCoV-19/England/MILK-A7CFDB/2020\|EPI_ISL_606329\|20 | .................... | .................... | .................... | .................... | .................... | .................... |
| O | 5655 hCoV-19/England/MILK-A7DAF7/2020\|EPI_ISL_606374\|20 | .................... | .................... | .................... | .................... | .................... | .................... |
| O | 5656 hCoV-19/England/ALDP-A6EFDC/2020\|EPI_ISL_607630\|20 | .................... | .................... | .................... | .................... | .................... | .................... |
| O | 5657 hCoV-19/England/ALDP-A6E482/2020\|EPI_ISL_607631\|20 | .................... | .................... | .................... | .................... | .................... | .................... |
| O | 5658 hCoV-19/England/ALDP-A6DE67/2020\|EPI_ISL_607632\|20 | .................... | .................... | .................... | .................... | .................... | .................... |
| O | 5659 hCoV-19/England/ALDP-A6E974/2020\|EPI_ISL_607633\|20 | .................... | .A.................. | .................... | .................... | .................... | ...A................ |
| O | 5660 hCoV-19/England/ALDP-A6D82D/2020\|EPI_ISL_607634\|20 | .................... | .................... | .................... | .................... | .................... | .................... |
| O | 5661 hCoV-19/England/ALDP-A6ECD5/2020\|EPI_ISL_607636\|20 | .................... | .................... | .................... | .................... | .................... | .................... |
| O | 5662 hCoV-19/England/ALDP-A6F3B1/2020\|EPI_ISL_607637\|20 | .................... | .A.................. | .................... | .................... | .................... | ...A................ |
| O | 5663 hCoV-19/England/ALDP-A6E1C7/2020\|EPI_ISL_607638\|20 | .................... | .................... | .................... | .................... | .................... | .................... |
| O | 5664 hCoV-19/England/ALDP-A6F9FB/2020\|EPI_ISL_607640\|20 | .................... | .................... | .................... | .................... | .................... | .................... |
| O | 5665 hCoV-19/England/ALDP-A70414/2020\|EPI_ISL_607641\|20 | .................... | .................... | .................... | .................... | .................... | .................... |
| O | 5666 hCoV-19/England/ALDP-A70371/2020\|EPI_ISL_607642\|20 | .................... | .................... | .................... | .................... | .................... | .................... |
| O | 5667 hCoV-19/England/ALDP-A6DF73/2020\|EPI_ISL_607644\|20 | .................... | .................... | .................... | .................... | .................... | .................... |
| O | 5668 hCoV-19/England/ALDP-A6F490/2020\|EPI_ISL_607645\|20 | .................... | .................... | .................... | .................... | .................... | .................... |
| O | 5669 hCoV-19/England/ALDP-A6F005/2020\|EPI_ISL_607646\|20 | .................... | .................... | .................... | .................... | .................... | .................... |
| O | 5670 hCoV-19/England/ALDP-A6DC4F/2020\|EPI_ISL_607647\|20 | .................... | .................... | .................... | .................... | .................... | .................... |
| O | 5671 hCoV-19/England/ALDP-A6E455/2020\|EPI_ISL_607648\|20 | .................... | .................... | .................... | .................... | .................... | .................... |
| O | 5672 hCoV-19/England/ALDP-A70775/2020\|EPI_ISL_607650\|20 | .................... | .................... | .................... | .................... | .................... | .................... |
| O | 5673 hCoV-19/England/ALDP-A6E18B/2020\|EPI_ISL_607652\|20 | .................... | .................... | .................... | .................... | .................... | .................... |
| O | 5674 hCoV-19/England/ALDP-A6E9FC/2020\|EPI_ISL_607655\|20 | .................... | .................... | .................... | .................... | .................... | .................... |
| O | 5675 hCoV-19/England/ALDP-A6EAAE/2020\|EPI_ISL_607657\|20 | .................... | .................... | .................... | .................... | .................... | .................... |
| O | 5676 hCoV-19/England/ALDP-A6E1B8/2020\|EPI_ISL_607658\|20 | .................... | .................... | .................... | .................... | .................... | .................... |
| O | 5677 hCoV-19/England/ALDP-A6F551/2020\|EPI_ISL_607659\|20 | .................... | .................... | .................... | .................... | .................... | .................... |
| O | 5678 hCoV-19/England/ALDP-A6900B/2020\|EPI_ISL_608900\|20 | .................... | .................... | .................... | .................... | .................... | .................... |

| | | | | | | | | |
|---|---|---|---|---|---|---|---|---|
| O | 5679 | hCoV-19/England/MILK-A6BFDF/2020\|EPI_ISL_608901\|20 | ···················· | ···················· | ···················· | ···················· | ···················· | ···················· |
| O | 5680 | hCoV-19/England/ALDP-A67D42/2020\|EPI_ISL_608903\|20 | ···················· | ·A·················· | ···················· | ···················· | ···················· | ···················· |
| O | 5681 | hCoV-19/England/ALDP-A682F7/2020\|EPI_ISL_608904\|20 | ···················· | ···················· | ···················· | ···················· | ···················· | ···················· |
| O | 5682 | hCoV-19/England/MILK-A6C123/2020\|EPI_ISL_608905\|20 | ···················· | ···················· | ···················· | ···················· | ···················· | ···················· |
| O | 5683 | hCoV-19/England/CAMC-A64A3F/2020\|EPI_ISL_608906\|20 | ···················· | ···················· | ···················· | ···················· | ···················· | ···················· |
| O | 5684 | hCoV-19/England/CAMC-A64853/2020\|EPI_ISL_608908\|20 | ···················· | ···················· | ···················· | ···················· | ···················· | ···················· |
| O | 5685 | hCoV-19/England/ALDP-A67D9D/2020\|EPI_ISL_608909\|20 | ···················· | ···················· | ···················· | ···················· | ···················· | ···················· |
| O | 5686 | hCoV-19/England/CAMC-A6462C/2020\|EPI_ISL_608910\|20 | ···················· | ···················· | ···················· | ···················· | ···················· | ···················· |
| O | 5687 | hCoV-19/England/ALDP-A68330/2020\|EPI_ISL_608911\|20 | ···················· | ···················· | ···················· | ···················· | ···················· | ···················· |
| O | 5688 | hCoV-19/England/CAMC-A647FC/2020\|EPI_ISL_608912\|20 | ···················· | ···················· | ···················· | ···················· | ···················· | ···················· |
| O | 5689 | hCoV-19/England/ALDP-A693C6/2020\|EPI_ISL_608913\|20 | ···················· | ···················· | ···················· | ···················· | ···················· | ···················· |
| O | 5690 | hCoV-19/England/ALDP-A680A2/2020\|EPI_ISL_608914\|20 | ···················· | ·A·················· | ···················· | ···················· | ···················· | ··A················· |
| O | 5691 | hCoV-19/England/ALDP-A6801B/2020\|EPI_ISL_608915\|20 | ···················· | ···················· | ···················· | ···················· | ···················· | ···················· |
| O | 5692 | hCoV-19/England/ALDP-A6933F/2020\|EPI_ISL_608916\|20 | ···················· | ···················· | ···················· | ···················· | ···················· | ···················· |
| O | 5693 | hCoV-19/England/ALDP-A68BDE/2020\|EPI_ISL_608917\|20 | ···················· | ···················· | ···················· | ···················· | ···················· | ···················· |
| O | 5694 | hCoV-19/England/CAMC-A64C66/2020\|EPI_ISL_608960\|20 | ···················· | ···················· | ···················· | ···················· | ···················· | ···················· |
| O | 5695 | hCoV-19/England/ALDP-A694E1/2020\|EPI_ISL_608961\|20 | ···················· | ···················· | ···················· | ···················· | ···················· | ···················· |
| O | 5696 | hCoV-19/England/ALDP-A69320/2020\|EPI_ISL_608963\|20 | ···················· | ···················· | ···················· | ···················· | ···················· | ···················· |
| O | 5697 | hCoV-19/England/ALDP-A69575/2020\|EPI_ISL_608964\|20 | ···················· | ···················· | ···················· | ···················· | ···················· | ···················· |
| O | 5698 | hCoV-19/England/CAMC-A64817/2020\|EPI_ISL_608965\|20 | ···················· | ···················· | ···················· | ···················· | ···················· | ···················· |
| O | 5699 | hCoV-19/England/ALDP-A69038/2020\|EPI_ISL_608966\|20 | ···················· | ···················· | ···················· | ···················· | ···················· | ···················· |
| O | 5700 | hCoV-19/England/CAMC-A64677/2020\|EPI_ISL_608968\|20 | ···················· | ·A·················· | ···················· | ···················· | ···················· | ··A················· |
| O | 5701 | hCoV-19/England/CAMC-A6497E/2020\|EPI_ISL_608969\|20 | ···················· | ···················· | ···················· | ···················· | ···················· | ···················· |
| O | 5702 | hCoV-19/England/ALDP-A68A86/2020\|EPI_ISL_608970\|20 | ···················· | ···················· | ···················· | ···················· | ···················· | ···················· |
| O | 5703 | hCoV-19/England/ALDP-A68A2C/2020\|EPI_ISL_608972\|20 | ···················· | ···················· | ···················· | ···················· | ···················· | ···················· |
| O | 5704 | hCoV-19/England/ALDP-A67D33/2020\|EPI_ISL_608973\|20 | ···················· | ···················· | ···················· | ···················· | ···················· | ···················· |
| O | 5705 | hCoV-19/England/CAMC-A64738/2020\|EPI_ISL_608974\|20 | ···················· | ···················· | ···················· | ···················· | ···················· | ···················· |
| O | 5706 | hCoV-19/England/MILK-A6BE69/2020\|EPI_ISL_608977\|20 | ···················· | ···················· | ···················· | ···················· | ···················· | ···················· |
| O | 5707 | hCoV-19/England/MILK-A6C0DB/2020\|EPI_ISL_608978\|20 | ···················· | ···················· | ···················· | ···················· | ···················· | ···················· |
| O | 5708 | hCoV-19/England/ALDP-A67FA6/2020\|EPI_ISL_608979\|20 | ···················· | ···················· | ···················· | ···················· | ···················· | ···················· |
| O | 5709 | hCoV-19/England/ALDP-A69056/2020\|EPI_ISL_608981\|20 | ···················· | ···················· | ···················· | ···················· | ···················· | ···················· |
| O | 5710 | hCoV-19/England/ALDP-A693B7/2020\|EPI_ISL_608983\|20 | ···················· | ···················· | ···················· | ···················· | ···················· | ···················· |
| O | 5711 | hCoV-19/England/ALDP-A68084/2020\|EPI_ISL_608984\|20 | ···················· | ···················· | ···················· | ···················· | ···················· | ···················· |
| O | 5712 | hCoV-19/England/ALDP-A67FE2/2020\|EPI_ISL_608985\|20 | ···················· | ·A··A··············· | ···················· | ···················· | ···················· | ··A················· |
| O | 5713 | hCoV-19/England/MILK-A6BD11/2020\|EPI_ISL_608986\|20 | ···················· | ···················· | ···················· | ···················· | ···················· | ···················· |
| O | 5714 | hCoV-19/England/CAMC-A64A99/2020\|EPI_ISL_608987\|20 | ···················· | ···················· | ···················· | ···················· | ·····T·············· | ···················· |
| O | 5715 | hCoV-19/England/ALDP-A69609/2020\|EPI_ISL_608988\|20 | ···················· | ···················· | ···················· | ···················· | ·····T·············· | ···················· |
| O | 5716 | hCoV-19/England/ALDP-A67D8E/2020\|EPI_ISL_608989\|20 | ···················· | ···················· | ···················· | ···················· | ···················· | ···················· |
| O | 5717 | hCoV-19/England/MILK-A191AD/2020\|EPI_ISL_612117\|20 | ···················· | ···················· | ···················· | ···················· | ···················· | ···················· |
| O | 5718 | hCoV-19/England/EXET-138165/2020\|EPI_ISL_612118\|20 | ···················· | ···················· | ···················· | ···················· | ···················· | ···················· |
| O | 5719 | hCoV-19/Wales/PHWC-4807B4/2020\|EPI_ISL_612119\|2020 | ···················· | ···················· | ···················· | ···················· | ···················· | ···················· |
| O | 5720 | hCoV-19/Wales/PHWC-48208A/2020\|EPI_ISL_612120\|2020 | ···················· | ···················· | ···················· | ···················· | ···················· | ···················· |
| O | 5721 | hCoV-19/England/MILK-A18D31/2020\|EPI_ISL_612121\|20 | ···················· | ···················· | ···················· | ···················· | ···················· | ···················· |
| O | 5722 | hCoV-19/England/CAMB-1B6C8C/2020\|EPI_ISL_612123\|20 | ···················· | ···················· | ···················· | ···················· | ···················· | ···················· |
| O | 5723 | hCoV-19/Scotland/CVR4697/2020\|EPI_ISL_612124\|2020- | ···················· | ···················· | ···················· | ···················· | ···················· | ···················· |
| O | 5724 | hCoV-19/Scotland/CVR4698/2020\|EPI_ISL_612125\|2020- | ···················· | ···················· | ···················· | ···················· | ···················· | ···················· |
| O | 5725 | hCoV-19/Scotland/EDB8337/2020\|EPI_ISL_612126\|2020- | ···················· | ···················· | ···················· | ···················· | ···················· | ···················· |
| O | 5726 | hCoV-19/Scotland/EDB8379/2020\|EPI_ISL_612127\|2020- | ···················· | ···················· | ···················· | ···················· | ···················· | ···················· |
| O | 5727 | hCoV-19/Scotland/EDB8450/2020\|EPI_ISL_612128\|2020- | ···················· | ···················· | ···················· | ···················· | ···················· | ···················· |
| O | 5728 | hCoV-19/Scotland/EDB8453/2020\|EPI_ISL_612129\|2020- | ···················· | ···················· | ···················· | ···················· | ···················· | ···················· |
| O | 5729 | hCoV-19/England/EXET-138253/2020\|EPI_ISL_612130\|20 | ···················· | ···················· | ···················· | ···················· | ···················· | ···················· |
| O | 5730 | hCoV-19/England/LOND-D9CC0/2020\|EPI_ISL_612131\|202 | ···················· | ···················· | ···················· | ···················· | ···················· | ···················· |
| O | 5731 | hCoV-19/England/MILK-A18D13/2020\|EPI_ISL_612132\|20 | ···················· | ···················· | ···················· | ···················· | ···················· | ···················· |
| O | 5732 | hCoV-19/England/MILK-A18D40/2020\|EPI_ISL_612133\|20 | ···················· | ···················· | ···················· | ···················· | ···················· | ···················· |
| O | 5733 | hCoV-19/England/MILK-A228CA/2020\|EPI_ISL_612134\|20 | ···················· | ···················· | ···················· | ···················· | ···················· | ···················· |
| O | 5734 | hCoV-19/Wales/PHWC-47EFFD/2020\|EPI_ISL_612135\|2020 | ···················· | ···················· | ···················· | ···················· | ···················· | ···················· |
| O | 5735 | hCoV-19/Wales/PHWC-47F2F3/2020\|EPI_ISL_612136\|2020 | ···················· | ···················· | ···················· | ···················· | ···················· | ···················· |
| O | 5736 | hCoV-19/Wales/PHWC-47F396/2020\|EPI_ISL_612137\|2020 | ···················· | ···················· | ···················· | ···················· | ···················· | ···················· |
| O | 5737 | hCoV-19/Wales/PHWC-47F3A5/2020\|EPI_ISL_612138\|2020 | ···················· | ···················· | ···················· | ···················· | ···················· | ···················· |
| O | 5738 | hCoV-19/Wales/PHWC-47F42A/2020\|EPI_ISL_612139\|2020 | ···················· | ···················· | ···················· | ···················· | ···················· | ···················· |
| O | 5739 | hCoV-19/Wales/PHWC-47F6BB/2020\|EPI_ISL_612140\|2020 | ···················· | ···················· | ···················· | ···················· | ···················· | ···················· |
| O | 5740 | hCoV-19/Wales/PHWC-47F93A/2020\|EPI_ISL_612141\|2020 | ···················· | ···················· | ···················· | ···················· | ···················· | ···················· |
| O | 5741 | hCoV-19/Wales/PHWC-47FAEC/2020\|EPI_ISL_612142\|2020 | ···················· | ···················· | ···················· | ···················· | ···················· | ···················· |
| O | 5742 | hCoV-19/Wales/PHWC-47FB43/2020\|EPI_ISL_612143\|2020 | ···················· | ···················· | ···················· | ···················· | ···················· | ···················· |
| O | 5743 | hCoV-19/Wales/PHWC-47FE95/2020\|EPI_ISL_612144\|2020 | ···················· | ···················· | ···················· | ···················· | ···················· | ···················· |
| O | 5744 | hCoV-19/Wales/PHWC-47FFB0/2020\|EPI_ISL_612145\|2020 | ···················· | ···················· | ···················· | ···················· | ···················· | ···················· |
| O | 5745 | hCoV-19/Wales/PHWC-4806A8/2020\|EPI_ISL_612146\|2020 | ···················· | ···················· | ···················· | ···················· | ···················· | ···················· |
| O | 5746 | hCoV-19/Wales/PHWC-480954/2020\|EPI_ISL_612147\|2020 | ···················· | ···················· | ···················· | ···················· | ···················· | ···················· |
| O | 5747 | hCoV-19/Wales/PHWC-4809BE/2020\|EPI_ISL_612148\|2020 | ···················· | ···················· | ···················· | ···················· | ···················· | ···················· |
| O | 5748 | hCoV-19/Wales/PHWC-480C97/2020\|EPI_ISL_612149\|2020 | ···················· | ···················· | ···················· | ···················· | ···················· | ···················· |
| O | 5749 | hCoV-19/Wales/PHWC-480E46/2020\|EPI_ISL_612150\|2020 | ···················· | ···················· | ···················· | ···················· | ···················· | ···················· |
| O | 5750 | hCoV-19/Wales/PHWC-480E82/2020\|EPI_ISL_612151\|2020 | ···················· | ···················· | ···················· | ···················· | ···················· | ···················· |
| O | 5751 | hCoV-19/Wales/PHWC-481021/2020\|EPI_ISL_612152\|2020 | ···················· | ···················· | ···················· | ···················· | ···················· | ···················· |
| O | 5752 | hCoV-19/Wales/PHWC-4811D3/2020\|EPI_ISL_612153\|2020 | ···················· | ···················· | ···················· | ···················· | ···················· | ···················· |
| O | 5753 | hCoV-19/Wales/PHWC-4816D4/2020\|EPI_ISL_612154\|2020 | ···················· | ···················· | ···················· | ···················· | ···················· | ···················· |
| O | 5754 | hCoV-19/Wales/PHWC-481ECD/2020\|EPI_ISL_612155\|2020 | ···················· | ···················· | ···················· | ···················· | ···················· | ···················· |
| O | 5755 | hCoV-19/Wales/PHWC-482020/2020\|EPI_ISL_612156\|2020 | ···················· | ···················· | ···················· | ···················· | ···················· | ···················· |
| O | 5757 | hCoV-19/Scotland/QEUH-A766FA/2020\|EPI_ISL_612158\|2 | ···················· | ···················· | ···················· | ···················· | ···················· | ···················· |
| O | 5759 | hCoV-19/Wales/PHWC-480A33/2020\|EPI_ISL_612204\|2020 | ···················· | ···················· | ···················· | ···················· | ···················· | ···················· |
| O | 5760 | hCoV-19/Wales/PHWC-480A51/2020\|EPI_ISL_612205\|2020 | ···················· | ···················· | ···················· | ···················· | ···················· | ···················· |

| | | | | | | | | | |
|---|---|---|---|---|---|---|---|---|---|
| O | 5761 | hCoV-19/Wales/PHWC-480D76/2020\|EPI_ISL_612206\|2020 | .................... | .................... | .................... | .................... | .................... | .................... | |
| O | 5762 | hCoV-19/England/BIRM-65AFC/2020\|EPI_ISL_612207\|202 | .................... | .................... | .................... | .................... | .................... | .................... | |
| O | 5764 | hCoV-19/England/BIRM-65B9F/2020\|EPI_ISL_612209\|202 | .................... | .................... | .................... | .................... | .................... | .................... | |
| O | 5768 | hCoV-19/England/BIRM-65C23/2020\|EPI_ISL_612213\|202 | .................... | .A.................. | .................... | .................... | .................... | ..A................. | |
| O | 5769 | hCoV-19/England/BIRM-65C50/2020\|EPI_ISL_612214\|202 | .................... | .................... | .................... | .................... | .................... | .................... | |
| O | 5770 | hCoV-19/England/BIRM-65C7E/2020\|EPI_ISL_612215\|202 | .................... | .................... | .................... | .................... | .................... | .................... | |
| O | 5771 | hCoV-19/England/BIRM-65C9C/2020\|EPI_ISL_612216\|202 | .................... | .................... | .................... | .................... | .................... | .................... | |
| O | 5772 | hCoV-19/England/BIRM-65CAB/2020\|EPI_ISL_612217\|202 | .................... | .................... | .................... | .................... | .................... | .................... | |
| O | 5773 | hCoV-19/Guadeloupe/IPG-7328/2020\|EPI_ISL_613428\|20 | .................... | .................... | .................... | .................... | .................... | .................... | |
| O | 5774 | hCoV-19/Saint Barthelemy/IPG-7329/2020\|EPI_ISL_613 | .................... | .................... | .................... | .................... | .................... | .................... | |
| O | 5775 | hCoV-19/Guadeloupe/IPG-7330/2020\|EPI_ISL_613430\|20 | .................... | .................... | .................... | .................... | .................... | .................... | |
| O | 5776 | hCoV-19/Guadeloupe/IPG-7331/2020\|EPI_ISL_613431\|20 | .................... | .................... | .................... | .................... | .................... | .................... | |
| O | 5777 | hCoV-19/Guadeloupe/IPG-7333/2020\|EPI_ISL_613432\|20 | .................... | .................... | .................... | .................... | .................... | .................... | |
| O | 5778 | hCoV-19/Guadeloupe/IPG-7334/2020\|EPI_ISL_613433\|20 | .................... | .................... | .................... | .................... | .................... | .................... | |
| O | 5779 | hCoV-19/Guadeloupe/IPG-7335/2020\|EPI_ISL_613434\|20 | .................... | .................... | .................... | .................... | .................... | .................... | |
| O | 5780 | hCoV-19/Guadeloupe/IPG-7336/2020\|EPI_ISL_613435\|20 | .................... | .................... | .................... | .................... | .................... | .................... | |
| O | 5781 | hCoV-19/Guadeloupe/IPG-7337/2020\|EPI_ISL_613436\|20 | .................... | .................... | .................... | .................... | .................... | .................... | |
| O | 5782 | hCoV-19/Guadeloupe/IPG-7338/2020\|EPI_ISL_613437\|20 | .................... | .................... | .................... | .................... | .................... | .................... | |
| O | 5783 | hCoV-19/Guadeloupe/IPG-7339/2020\|EPI_ISL_613438\|20 | .................... | .................... | .................... | .................... | .................... | .................... | |
| O | 5784 | hCoV-19/Guadeloupe/IPG-7340/2020\|EPI_ISL_613439\|20 | .................... | .................... | .................... | .................... | .................... | .................... | |
| O | 5785 | hCoV-19/Guadeloupe/IPG-7341/2020\|EPI_ISL_613440\|20 | .................... | .................... | .................... | .................... | .................... | .................... | |
| O | 5786 | hCoV-19/Guadeloupe/IPG-7342/2020\|EPI_ISL_613441\|20 | .................... | .................... | .................... | .................... | .................... | .................... | |
| O | 5787 | hCoV-19/Guadeloupe/IPG-7344/2020\|EPI_ISL_613442\|20 | .................... | .................... | .................... | .................... | .................... | .................... | |
| O | 5788 | hCoV-19/Guadeloupe/IPG-7346/2020\|EPI_ISL_613443\|20 | .................... | .................... | .................... | .................... | .................... | .................... | |
| O | 5789 | hCoV-19/Guadeloupe/IPG-7347/2020\|EPI_ISL_613444\|20 | .................... | .................... | .................... | .................... | .................... | .................... | |
| O | 5790 | hCoV-19/Guadeloupe/IPG-7353/2020\|EPI_ISL_613445\|20 | .................... | .................... | .................... | .................... | .................... | .................... | |
| O | 5791 | hCoV-19/Saint Barthelemy/IPG-7358/2020\|EPI_ISL_613 | .................... | .................... | .................... | .................... | .................... | .................... | |
| O | 5792 | hCoV-19/Saint Martin/IPG-7343/2020\|EPI_ISL_613447\| | .................... | .................... | .................... | .................... | .................... | .................... | |
| O | 5793 | hCoV-19/Saint Martin/IPG-7348/2020\|EPI_ISL_613448\| | .................... | .................... | .................... | .................... | .................... | .................... | |
| O | 5794 | hCoV-19/Saint Martin/IPG-7349/2020\|EPI_ISL_613449\| | .................... | .................... | .................... | .................... | .................... | .................... | |
| O | 5795 | hCoV-19/Saint Martin/IPG-7350/2020\|EPI_ISL_613450\| | .................... | .................... | .................... | .................... | .................... | .................... | |
| O | 5796 | hCoV-19/Saint Martin/IPG-7351/2020\|EPI_ISL_613451\| | .................... | .................... | .................... | .................... | .................... | .................... | |
| O | 5797 | hCoV-19/Saint Martin/IPG-7352/2020\|EPI_ISL_613452\| | .................... | .................... | .................... | .................... | .................... | .................... | |
| O | 5798 | hCoV-19/Saint Martin/IPG-7356/2020\|EPI_ISL_613455\| | .................... | .................... | .................... | .................... | .................... | .................... | |
| O | 5799 | hCoV-19/Saint Martin/IPG-7357/2020\|EPI_ISL_613456\| | .................... | .................... | .................... | .................... | .................... | .................... | |
| O | 5800 | hCoV-19/Hong Kong/VM20106394/2020\|EPI_ISL_613457\|2 | .................... | .................... | .................... | .................... | .................... | .................... | |
| O | 5801 | hCoV-19/Turkey/KOC-IST-SP3/2020\|EPI_ISL_613459\|202 | .................... | .................... | .................... | .................... | .................... | .................... | |
| O | 5802 | hCoV-19/Turkey/KOC-IST-OD5/2020\|EPI_ISL_613460\|202 | .................... | .................... | .................... | .................... | .................... | .................... | |
| O | 5803 | hCoV-19/Turkey/KOC-IST-B288/2020\|EPI_ISL_613461\|20 | .................... | .................... | .................... | .................... | .................... | .................... | |
| O | 5804 | hCoV-19/Turkey/KOC-IST-B316/2020\|EPI_ISL_613462\|20 | .................... | .................... | .................... | .................... | .................... | .................... | |
| O | 5805 | hCoV-19/USA/MN-MDH-1963/2020\|EPI_ISL_613463\|2020-0 | .................... | .............A..... | .................... | .................... | .................... | .................... | |
| O | 5806 | hCoV-19/USA/MN-MDH-1962/2020\|EPI_ISL_613464\|2020-0 | .................... | .................... | .................... | .................... | .................... | .................... | |
| O | 5807 | hCoV-19/USA/MN-MDH-1961/2020\|EPI_ISL_613465\|2020-0 | .................... | .............A..... | .................... | .................... | .................... | .................... | |
| O | 5808 | hCoV-19/USA/MN-MDH-1960/2020\|EPI_ISL_613466\|2020-0 | .................... | .............A..... | .................... | .................... | .................... | .................... | |
| O | 5809 | hCoV-19/USA/MN-MDH-1959/2020\|EPI_ISL_613467\|2020-0 | .................... | .............A..... | .................... | .................... | .................... | .................... | |
| O | 5810 | hCoV-19/USA/MN-MDH-1958/2020\|EPI_ISL_613468\|2020-0 | .................... | .............A..... | .................... | .................... | .................... | .................... | |
| O | 5811 | hCoV-19/USA/MN-MDH-1957/2020\|EPI_ISL_613469\|2020-0 | .................... | .............A..... | .................... | .................... | .................... | .................... | |
| O | 5812 | hCoV-19/USA/MN-MDH-1956/2020\|EPI_ISL_613470\|2020-0 | .................... | .............A..... | .................... | .................... | .................... | .................... | |
| O | 5813 | hCoV-19/USA/MN-MDH-1955/2020\|EPI_ISL_613471\|2020-0 | .................... | .............A..... | .................... | .................... | .................... | .................... | |
| O | 5814 | hCoV-19/USA/MN-MDH-1954/2020\|EPI_ISL_613472\|2020-0 | .................... | .............A..... | .................... | .................... | .................... | .................... | |
| O | 5815 | hCoV-19/USA/MN-MDH-1953/2020\|EPI_ISL_613473\|2020-0 | .................... | .............A..... | .................... | .................... | .................... | .................... | |
| O | 5816 | hCoV-19/USA/MN-MDH-1952/2020\|EPI_ISL_613474\|2020-0 | .................... | .............A..... | .................... | .................... | .................... | .................... | |
| O | 5817 | hCoV-19/USA/MN-MDH-1951/2020\|EPI_ISL_613475\|2020-0 | .................... | .A.................. | .................... | .................... | .................... | .................... | |
| O | 5818 | hCoV-19/USA/MN-MDH-1950/2020\|EPI_ISL_613476\|2020-0 | .................... | .............A..... | .................... | .................... | .NNN................ | .................... | 1 |
| O | 5819 | hCoV-19/USA/MN-MDH-1949/2020\|EPI_ISL_613477\|2020-0 | .................... | .............A..... | .................... | .................... | .................... | .................... | |
| O | 5820 | hCoV-19/USA/MN-MDH-1948/2020\|EPI_ISL_613478\|2020-0 | .................... | .................... | .................... | .................... | .................... | .................... | |
| O | 5821 | hCoV-19/USA/MN-MDH-1947/2020\|EPI_ISL_613479\|2020-0 | .................... | .............A..... | .................... | .................... | .................... | .................... | |
| O | 5822 | hCoV-19/USA/MN-MDH-1946/2020\|EPI_ISL_613480\|2020-0 | .................... | .............A..... | .................... | .................... | .................... | .................... | |
| O | 5823 | hCoV-19/USA/MN-MDH-1945/2020\|EPI_ISL_613481\|2020-0 | .................... | .................... | .................... | .................... | .................... | .................... | |
| O | 5824 | hCoV-19/USA/MN-MDH-1944/2020\|EPI_ISL_613482\|2020-0 | .................... | .................... | .................... | .................... | .................... | .................... | |
| O | 5825 | hCoV-19/USA/MN-MDH-1943/2020\|EPI_ISL_613483\|2020-0 | .................... | .A.................. | .................... | .................... | .................... | .................... | |
| O | 5826 | hCoV-19/USA/MN-MDH-1942/2020\|EPI_ISL_613484\|2020-0 | .................... | .................... | .................... | .................... | .................... | .................... | |
| O | 5827 | hCoV-19/USA/MN-MDH-1941/2020\|EPI_ISL_613485\|2020-0 | .................... | .................... | .................... | .................... | .................... | .................... | |
| O | 5828 | hCoV-19/USA/MN-MDH-1940/2020\|EPI_ISL_613486\|2020-0 | .................... | .A.................. | .................... | .................... | .................... | .................... | |
| O | 5829 | hCoV-19/USA/MN-MDH-1939/2020\|EPI_ISL_613487\|2020-0 | .................... | ......A......A..... | .................... | .................... | .................... | .................... | |
| O | 5830 | hCoV-19/USA/MN-MDH-1938/2020\|EPI_ISL_613488\|2020-0 | .................... | .................... | .................... | .................... | .................... | .................... | |
| O | 5831 | hCoV-19/USA/MN-MDH-1937/2020\|EPI_ISL_613489\|2020-0 | .................... | .................... | .................... | .................... | .................... | .................... | |
| O | 5832 | hCoV-19/USA/MN-MDH-1936/2020\|EPI_ISL_613490\|2020-0 | .................... | .................... | .................... | .................... | .................... | .................... | |
| O | 5833 | hCoV-19/USA/MN-MDH-1935/2020\|EPI_ISL_613491\|2020-1 | .................... | .................... | .................... | .................... | .................... | .................... | |
| O | 5834 | hCoV-19/USA/MN-MDH-1934/2020\|EPI_ISL_613492\|2020-1 | .................... | .................... | .................... | .................... | .................... | .................... | |
| O | 5835 | hCoV-19/USA/MN-MDH-1933/2020\|EPI_ISL_613493\|2020-1 | .................... | .................... | .................... | .................... | .................... | .................... | |
| O | 5836 | hCoV-19/USA/MN-MDH-1932/2020\|EPI_ISL_613494\|2020-1 | .................... | .................... | .................... | .................... | .................... | .................... | |
| O | 5837 | hCoV-19/USA/MN-MDH-1931/2020\|EPI_ISL_613495\|2020-1 | .................... | .................... | .................... | .................... | .................... | .................... | |
| O | 5838 | hCoV-19/USA/MN-MDH-1930/2020\|EPI_ISL_613496\|2020-1 | .................... | .................... | .................... | .................... | .................... | .................... | |
| O | 5839 | hCoV-19/USA/MN-MDH-1929/2020\|EPI_ISL_613497\|2020-1 | .................... | .................... | .................... | .................... | .................... | .................... | |
| O | 5840 | hCoV-19/USA/MN-MDH-1928/2020\|EPI_ISL_613498\|2020-1 | .................... | .................... | .................... | .................... | .................... | .................... | |
| O | 5841 | hCoV-19/USA/MN-MDH-1927/2020\|EPI_ISL_613499\|2020-1 | .................... | .................... | .................... | .................... | .................... | .................... | |
| O | 5842 | hCoV-19/USA/MN-MDH-1926/2020\|EPI_ISL_613500\|2020-1 | .................... | .................... | .................... | .................... | .................... | .................... | |
| O | 5843 | hCoV-19/USA/MN-MDH-1925/2020\|EPI_ISL_613501\|2020-1 | .................... | .................... | .................... | .................... | ..NN................ | .................... | |
| O | 5844 | hCoV-19/USA/MN-MDH-1924/2020\|EPI_ISL_613502\|2020-1 | .................... | .................... | .................... | .................... | .................... | .................... | |

| | | | | | | | | | |
|---|---|---|---|---|---|---|---|---|---|
| O | 5845 | hCoV-19/USA/MN-MDH-1923/2020\|EPI_ISL_613503\|2020-1 | .................... | .................... | ..................... | .................... | .................... | .................... | |
| O | 5846 | hCoV-19/USA/MN-MDH-1922/2020\|EPI_ISL_613504\|2020-1 | .................... | .................... | ..................... | .................... | .................... | .................... | |
| O | 5847 | hCoV-19/USA/MN-MDH-1921/2020\|EPI_ISL_613505\|2020-1 | .................... | .................... | ..................... | .................... | .................... | .................... | |
| O | 5848 | hCoV-19/USA/MN-MDH-1920/2020\|EPI_ISL_613506\|2020-1 | .................... | .................... | ..................... | .................... | .................... | .................... | |
| O | 5849 | hCoV-19/USA/MN-MDH-1919/2020\|EPI_ISL_613507\|2020-1 | .................... | .................... | ..................... | .................... | .................... | .................... | |
| O | 5850 | hCoV-19/USA/MN-MDH-1918/2020\|EPI_ISL_613508\|2020-1 | .................... | .................... | ..................... | .................... | .................... | .................... | |
| O | 5851 | hCoV-19/USA/MN-MDH-1917/2020\|EPI_ISL_613509\|2020-1 | .................... | .................... | ..................... | .................... | .................... | .................... | |
| O | 5852 | hCoV-19/USA/MN-MDH-1916/2020\|EPI_ISL_613510\|2020-1 | .................... | .................... | ..................... | .................... | .................... | .................... | |
| O | 5853 | hCoV-19/USA/MN-MDH-1915/2020\|EPI_ISL_613511\|2020-1 | .................... | .................... | ..................... | .................... | .................... | .................... | |
| O | 5854 | hCoV-19/USA/MN-MDH-1914/2020\|EPI_ISL_613512\|2020-1 | .................... | .............A...... | ..................... | .................... | .................... | .................... | |
| O | 5855 | hCoV-19/USA/MN-MDH-1913/2020\|EPI_ISL_613513\|2020-1 | .................... | .............A...... | ..................... | .................... | .................... | .................... | |
| O | 5856 | hCoV-19/USA/MN-MDH-1912/2020\|EPI_ISL_613514\|2020-1 | .................... | .............A...... | ..................... | .................... | .................... | .................... | |
| O | 5857 | hCoV-19/USA/MN-MDH-1911/2020\|EPI_ISL_613515\|2020-1 | .................... | .............A...... | ..................... | .................... | .................... | .................... | |
| O | 5858 | hCoV-19/USA/MN-MDH-1910/2020\|EPI_ISL_613516\|2020-1 | .................... | .............A...... | ..................... | .................... | .................... | .................... | |
| O | 5859 | hCoV-19/USA/MN-MDH-1909/2020\|EPI_ISL_613517\|2020-1 | .................... | .............A...... | ..................... | .................... | .................... | .................... | |
| O | 5860 | hCoV-19/USA/MN-MDH-1908/2020\|EPI_ISL_613518\|2020-1 | .................... | .............A...... | ..................... | .................... | .................... | .................... | |
| O | 5861 | hCoV-19/USA/MN-MDH-1907/2020\|EPI_ISL_613519\|2020-1 | .................... | .............A...... | ..................... | .................... | .................... | .................... | |
| O | 5862 | hCoV-19/USA/MN-MDH-1906/2020\|EPI_ISL_613520\|2020-1 | .................... | .............A...... | ..................... | .................... | .................... | .................... | |
| O | 5863 | hCoV-19/USA/MN-MDH-1905/2020\|EPI_ISL_613521\|2020-1 | .................... | .............A...... | ..................... | .................... | .................... | .................... | |
| O | 5864 | hCoV-19/USA/MN-MDH-1904/2020\|EPI_ISL_613522\|2020-1 | .................... | .............A...... | ..................... | .................... | .................... | .................... | |
| O | 5865 | hCoV-19/USA/MN-MDH-1903/2020\|EPI_ISL_613523\|2020-1 | .................... | .................... | ..................... | .................... | .................... | .................... | |
| O | 5866 | hCoV-19/USA/MN-MDH-1902/2020\|EPI_ISL_613524\|2020-1 | .................... | .................... | ..................... | .................... | .................... | .................... | |
| O | 5867 | hCoV-19/USA/MN-MDH-1901/2020\|EPI_ISL_613525\|2020-1 | .................... | .................... | ..................... | .................... | .................... | .................... | |
| O | 5868 | hCoV-19/USA/MN-MDH-1899/2020\|EPI_ISL_613527\|2020-1 | .................... | .................... | ..................... | .................... | .................... | .................... | |
| O | 5869 | hCoV-19/USA/MN-MDH-1898/2020\|EPI_ISL_613528\|2020-1 | .................... | .................... | ..................... | .................... | .................... | .................... | |
| O | 5870 | hCoV-19/USA/MN-MDH-1897/2020\|EPI_ISL_613529\|2020-1 | .................... | .............A...... | ..................... | .................... | .................... | .................... | |
| O | 5871 | hCoV-19/USA/MN-MDH-1896/2020\|EPI_ISL_613530\|2020-1 | .................... | .................... | ..................... | .................... | .................... | .................... | |
| O | 5872 | hCoV-19/USA/MN-MDH-1895/2020\|EPI_ISL_613531\|2020-1 | .................... | .................... | ..................... | .................... | .................... | .................... | |
| O | 5873 | hCoV-19/USA/MN-MDH-1894/2020\|EPI_ISL_613532\|2020-1 | .................... | .A.................. | ..................... | .................... | .................... | .................... | |
| O | 5874 | hCoV-19/USA/MN-MDH-1893/2020\|EPI_ISL_613533\|2020-1 | .................... | .............A...... | ..................... | .................... | .................... | .................... | |
| O | 5875 | hCoV-19/USA/MN-MDH-1892/2020\|EPI_ISL_613534\|2020-1 | .................... | .............A...... | ..................... | .................... | .................... | .................... | |
| O | 5876 | hCoV-19/USA/MN-MDH-1891/2020\|EPI_ISL_613535\|2020-1 | .................... | .............A...... | ..................... | .................... | .................... | .................... | |
| O | 5877 | hCoV-19/USA/MN-MDH-1890/2020\|EPI_ISL_613536\|2020-1 | .................... | .............A...... | ..................... | .................... | .................... | .................... | |
| O | 5878 | hCoV-19/USA/MN-MDH-1889/2020\|EPI_ISL_613537\|2020-1 | .................... | .............A...... | ..................... | .................... | .................... | .................... | |
| O | 5879 | hCoV-19/USA/MN-MDH-1888/2020\|EPI_ISL_613538\|2020-1 | .................... | .............A...... | ..................... | .................... | NNN................. | .................... | 1 |
| O | 5880 | hCoV-19/USA/MN-MDH-1887/2020\|EPI_ISL_613539\|2020-1 | .................... | .............A...... | ..................... | .................... | .................... | .................... | |
| O | 5881 | hCoV-19/USA/MN-MDH-1886/2020\|EPI_ISL_613540\|2020-1 | .................... | .............A...... | ..................... | .................... | .................... | .................... | |
| O | 5882 | hCoV-19/Denmark/DCGC-746/2020\|EPI_ISL_613541\|2020- | .................... | .................... | ..................... | .................... | .................... | .................... | |
| O | 5883 | hCoV-19/Denmark/DCGC-751/2020\|EPI_ISL_613542\|2020- | .................... | .................... | ..................... | .................... | .................... | .................... | |
| O | 5884 | hCoV-19/Denmark/DCGC-753/2020\|EPI_ISL_613543\|2020- | .................... | .................... | ..................... | .................... | .................... | .................... | |
| O | 5885 | hCoV-19/France/BRE-BR9055/2020\|EPI_ISL_613545\|2020 | .................... | .................... | ..................... | .................... | .................... | .................... | |
| O | 5887 | hCoV-19/France/BRE-BR9057/2020\|EPI_ISL_613547\|2020 | .................... | .................... | ..................... | .................... | .................... | ...............A.... | |
| O | 5888 | hCoV-19/France/BRE-BR9059/2020\|EPI_ISL_613548\|2020 | .................... | .................... | ..................... | .................... | .................... | .................... | |
| O | 5889 | hCoV-19/England/ALDP-A87ADC/2020\|EPI_ISL_606222\|20 | .................... | .................... | ..................... | .................... | .................... | .................... | |
| O | 5890 | hCoV-19/England/ALDP-A87BF7/2020\|EPI_ISL_606223\|20 | .................... | .................... | ..................... | .................... | .................... | .................... | |
| O | 5891 | hCoV-19/England/ALDP-A86EC2/2020\|EPI_ISL_606224\|20 | .................... | .................... | ..................... | .................... | .................... | .................... | |
| O | 5892 | hCoV-19/England/ALDP-A86EB3/2020\|EPI_ISL_606225\|20 | .................... | .................... | ..................... | .................... | .................... | .................... | |
| O | 5893 | hCoV-19/England/ALDP-A86484/2020\|EPI_ISL_606227\|20 | .................... | .A.................. | ..................... | .................... | .................... | .................... | |
| O | 5894 | hCoV-19/England/ALDP-A8790C/2020\|EPI_ISL_606228\|20 | .................... | .................... | ..................... | .................... | .................... | .................... | |
| O | 5895 | hCoV-19/England/ALDP-A863D2/2020\|EPI_ISL_606229\|20 | .................... | .................... | ..................... | .................... | .................... | .................... | |
| O | 5896 | hCoV-19/England/ALDP-A87878/2020\|EPI_ISL_606230\|20 | .................... | .................... | ..................... | .................... | .................... | .................... | |
| O | 5897 | hCoV-19/England/ALDP-A872B6/2020\|EPI_ISL_606231\|20 | .................... | .................... | ..................... | .................... | .................... | .................... | |
| O | 5898 | hCoV-19/England/ALDP-A861BA/2020\|EPI_ISL_606232\|20 | .................... | .................... | ..................... | .................... | .................... | .................... | |
| O | 5899 | hCoV-19/England/ALDP-A865AF/2020\|EPI_ISL_606233\|20 | .................... | .................... | ..................... | .................... | .................... | .................... | |
| O | 5900 | hCoV-19/England/ALDP-A86633/2020\|EPI_ISL_606234\|20 | .................... | .................... | ..................... | .................... | .................... | .................... | |
| O | 5901 | hCoV-19/England/ALDP-A86CAA/2020\|EPI_ISL_606235\|20 | .................... | .................... | ..................... | .................... | .................... | .................... | |
| O | 5902 | hCoV-19/England/ALDP-A86CF5/2020\|EPI_ISL_606236\|20 | .................... | .................... | ..................... | .................... | .................... | .................... | |
| O | 5903 | hCoV-19/England/ALDP-A873A4/2020\|EPI_ISL_606237\|20 | .................... | .................... | ..................... | .................... | .................... | .................... | |
| O | 5904 | hCoV-19/England/ALDP-A878F0/2020\|EPI_ISL_606238\|20 | .................... | .................... | ..................... | .................... | .................... | .................... | |
| O | 5905 | hCoV-19/England/ALDP-A871AA/2020\|EPI_ISL_606239\|20 | .................... | .................... | ..................... | .................... | .................... | .................... | |
| O | 5906 | hCoV-19/England/ALDP-A86439/2020\|EPI_ISL_606240\|20 | .................... | .................... | ..................... | .................... | .................... | .................... | |
| O | 5907 | hCoV-19/England/ALDP-A86554/2020\|EPI_ISL_606241\|20 | .................... | .................... | ..................... | .................... | .................... | .................... | |
| O | 5908 | hCoV-19/England/ALDP-A874B0/2020\|EPI_ISL_606243\|20 | .................... | .................... | ..................... | .................... | .................... | .................... | |
| O | 5909 | hCoV-19/England/ALDP-A87B60/2020\|EPI_ISL_606244\|20 | .................... | .A.................. | ..................... | .................... | .................... | .................... | |
| O | 5910 | hCoV-19/England/ALDP-A8708F/2020\|EPI_ISL_606245\|20 | .................... | .................... | ..................... | .................... | .................... | .................... | |
| O | 5911 | hCoV-19/England/ALDP-A86FDE/2020\|EPI_ISL_606246\|20 | .................... | .................... | ..................... | .................... | .................... | .................... | |
| O | 5912 | hCoV-19/England/ALDP-A8618D/2020\|EPI_ISL_606248\|20 | .................... | .................... | ..................... | .................... | .................... | .................... | |
| O | 5913 | hCoV-19/England/ALDP-A87641/2020\|EPI_ISL_606249\|20 | .................... | .................... | ..................... | .................... | .................... | .................... | |
| O | 5914 | hCoV-19/England/ALDP-A86080/2020\|EPI_ISL_606250\|20 | .................... | .................... | ..................... | .................... | .................... | .................... | |
| O | 5915 | hCoV-19/England/ALDP-A875CC/2020\|EPI_ISL_606251\|20 | .................... | .................... | ..................... | .................... | .................... | .................... | |
| O | 5916 | hCoV-19/England/ALDP-A8723E/2020\|EPI_ISL_606252\|20 | .................... | .................... | ..................... | .................... | .................... | .................... | |
| O | 5917 | hCoV-19/England/ALDP-A87BCA/2020\|EPI_ISL_606253\|20 | .................... | .................... | ..................... | .................... | .................... | .................... | |
| O | 5918 | hCoV-19/England/ALDP-A879C0/2020\|EPI_ISL_606254\|20 | .................... | .................... | ..................... | .................... | .................... | .................... | |
| O | 5919 | hCoV-19/England/ALDP-A872A7/2020\|EPI_ISL_606255\|20 | .................... | .................... | ..................... | .................... | .................... | .................... | |
| O | 5920 | hCoV-19/England/ALDP-A86642/2020\|EPI_ISL_606256\|20 | .................... | .................... | ..................... | .................... | .................... | .................... | |
| O | 5921 | hCoV-19/England/ALDP-A8777B/2020\|EPI_ISL_606257\|20 | .................... | .................... | ..................... | .................... | .................... | .................... | |
| O | 5922 | hCoV-19/England/ALDP-A8630F/2020\|EPI_ISL_606258\|20 | .................... | .................... | ..................... | .................... | .................... | .................... | |
| O | 5923 | hCoV-19/England/ALDP-A87623/2020\|EPI_ISL_606260\|20 | .................... | .................... | ..................... | .................... | .................... | .................... | |
| O | 5924 | hCoV-19/England/ALDP-A8624E/2020\|EPI_ISL_606263\|20 | .................... | .................... | ..................... | .................... | .................... | .................... | |
| O | 5925 | hCoV-19/England/ALDP-A862C6/2020\|EPI_ISL_606264\|20 | .................... | .................... | ..................... | .................... | .................... | .................... | |

| | | | | | | | |
|---|---|---|---|---|---|---|---|
| O | 5926 hCoV-19/England/ALDP-A86202/2020\|EPI_ISL_606265\|20 | ···················· | ···················· | ···················· | ···················· | ···················· | ···················· |
| O | 5927 hCoV-19/England/ALDP-A87702/2020\|EPI_ISL_606266\|20 | ···················· | ···················· | ···················· | ···················· | ···················· | ···················· |
| O | 5928 hCoV-19/England/ALDP-A87052/2020\|EPI_ISL_606268\|20 | ···················· | ···················· | ···················· | ···················· | ···················· | ···················· |
| O | 5929 hCoV-19/England/ALDP-A862E4/2020\|EPI_ISL_606269\|20 | ···················· | ···················· | ···················· | ···················· | ···················· | ···················· |
| O | 5930 hCoV-19/England/ALDP-A872C5/2020\|EPI_ISL_606270\|20 | ···················· | ···················· | ···················· | ···················· | ···················· | ···················· |
| O | 5931 hCoV-19/England/ALDP-A87140/2020\|EPI_ISL_606271\|20 | ···················· | ···················· | ···················· | ···················· | ···················· | ···················· |
| O | 5932 hCoV-19/England/ALDP-A8785A/2020\|EPI_ISL_606272\|20 | ···················· | ···················· | ···················· | ···················· | ···················· | ···················· |
| O | 5933 hCoV-19/England/ALDP-A8781E/2020\|EPI_ISL_606273\|20 | ···················· | ···················· | ···················· | ···················· | ···················· | ···················· |
| O | 5934 hCoV-19/England/ALDP-A87AEB/2020\|EPI_ISL_606274\|20 | ···················· | ···················· | ···················· | ···················· | ···················· | ···················· |
| O | 5935 hCoV-19/England/ALDP-A87A45/2020\|EPI_ISL_606275\|20 | ···················· | ···················· | ···················· | ···················· | ···················· | ···················· |
| O | 5936 hCoV-19/England/ALDP-A876C9/2020\|EPI_ISL_606276\|20 | ···················· | ···················· | ···················· | ···················· | ···················· | ···················· |
| O | 5937 hCoV-19/England/ALDP-A87C12/2020\|EPI_ISL_606277\|20 | ···················· | ···················· | ···················· | ···················· | ···················· | ···················· |
| O | 5938 hCoV-19/England/ALDP-A8724D/2020\|EPI_ISL_606278\|20 | ···················· | ···················· | ···················· | ···················· | ···················· | ···················· |
| O | 5939 hCoV-19/England/ALDP-A87395/2020\|EPI_ISL_606279\|20 | ···················· | ···················· | ···················· | ···················· | ······T············· | ···················· |
| O | 5940 hCoV-19/England/ALDP-A8631E/2020\|EPI_ISL_606280\|20 | ···················· | ···················· | ···················· | ···················· | ···················· | ···················· |
| O | 5941 hCoV-19/England/ALDP-A87896/2020\|EPI_ISL_606281\|20 | ···················· | ···················· | ···················· | ···················· | ···················· | ···················· |
| O | 5942 hCoV-19/England/ALDP-A878E1/2020\|EPI_ISL_606282\|20 | ···················· | ···················· | ···················· | ···················· | ···················· | ···················· |
| O | 5943 hCoV-19/England/ALDP-A872F2/2020\|EPI_ISL_606283\|20 | ···················· | ···················· | ···················· | ···················· | ···················· | ···················· |
| O | 5944 hCoV-19/England/ALDP-A865EB/2020\|EPI_ISL_606284\|20 | ···················· | ···················· | ···················· | ···················· | ···················· | ···················· |
| O | 5945 hCoV-19/England/ALDP-A87BD9/2020\|EPI_ISL_606285\|20 | ···················· | ···················· | ···················· | ···················· | ···················· | ···················· |
| O | 5946 hCoV-19/England/ALDP-A8616F/2020\|EPI_ISL_606287\|20 | ···················· | ···················· | ···················· | ···················· | ···················· | ···················· |
| O | 5947 hCoV-19/England/ALDP-A8609F/2020\|EPI_ISL_606288\|20 | ···················· | ···················· | ···················· | ···················· | ···················· | ···················· |
| O | 5948 hCoV-19/England/ALDP-A87966/2020\|EPI_ISL_606289\|20 | ···················· | ···················· | ···················· | ···················· | ···················· | ···················· |
| O | 5949 hCoV-19/England/ALDP-A875BD/2020\|EPI_ISL_606290\|20 | ···················· | ···················· | ···················· | ···················· | ···················· | ···················· |
| O | 5950 hCoV-19/England/ALDP-A87AAF/2020\|EPI_ISL_606292\|20 | ···················· | ···················· | ···················· | ···················· | ···················· | ···················· |
| O | 5951 hCoV-19/England/ALDP-A877E4/2020\|EPI_ISL_606293\|20 | ···················· | ···················· | ···················· | ···················· | ···················· | ···················· |
| O | 5952 hCoV-19/England/ALDP-A87A63/2020\|EPI_ISL_606294\|20 | ···················· | ···················· | ···················· | ···················· | ···················· | ···················· |
| O | 5953 hCoV-19/England/ALDP-A86475/2020\|EPI_ISL_606295\|20 | ···················· | ···················· | ···················· | ···················· | ···················· | ···················· |
| O | 5954 hCoV-19/England/MILK-A7D65D/2020\|EPI_ISL_606296\|20 | ···················· | ···················· | ···················· | ···················· | ···················· | ···················· |
| O | 5955 hCoV-19/England/MILK-A7D5BA/2020\|EPI_ISL_606297\|20 | ···················· | ···················· | ···················· | ···················· | ···················· | ···················· |
| O | 5956 hCoV-19/England/MILK-A7A74E/2020\|EPI_ISL_606298\|20 | ···················· | ···················· | ···················· | ···················· | ···················· | ···················· |
| O | 5957 hCoV-19/England/MILK-A7AD88/2020\|EPI_ISL_606299\|20 | ···················· | ···················· | ···················· | ···················· | ···················· | ···················· |
| O | 5958 hCoV-19/England/MILK-A7A67E/2020\|EPI_ISL_606320\|20 | ···················· | ···················· | ···················· | ···················· | ······T············· | ···············A···· |
| O | 5959 hCoV-19/England/MILK-A7CA16/2020\|EPI_ISL_606330\|20 | ···················· | ···················· | ···················· | ···················· | ···················· | ···················· |
| O | 5960 hCoV-19/England/MILK-A7AAAF/2020\|EPI_ISL_606331\|20 | ···················· | ···················· | ···················· | ···················· | ···················· | ···················· |
| O | 5961 hCoV-19/England/MILK-A7D8DF/2020\|EPI_ISL_606332\|20 | ···················· | ···················· | ···················· | ···················· | ···················· | ···················· |
| O | 5962 hCoV-19/England/MILK-A7DD67/2020\|EPI_ISL_606334\|20 | ···················· | ···················· | ···················· | ···················· | ···················· | ···················· |
| O | 5963 hCoV-19/England/MILK-A7CD59/2020\|EPI_ISL_606335\|20 | ···················· | ·A·················· | ···················· | ···················· | ···················· | ···A················ |
| O | 5964 hCoV-19/England/MILK-A7D6D5/2020\|EPI_ISL_606336\|20 | ···················· | ···················· | ···················· | ···················· | ···················· | ···················· |
| O | 5965 hCoV-19/England/MILK-A7D356/2020\|EPI_ISL_606337\|20 | ···················· | ···················· | ···················· | ···················· | ···················· | ···················· |
| O | 5966 hCoV-19/England/MILK-A7C0BA/2020\|EPI_ISL_606338\|20 | ···················· | ···················· | ···················· | ···················· | ···················· | ···················· |
| O | 5967 hCoV-19/England/MILK-A7CA61/2020\|EPI_ISL_606339\|20 | ···················· | ···················· | ···················· | ···················· | ···················· | ···················· |
| O | 5968 hCoV-19/England/MILK-A7B525/2020\|EPI_ISL_607551\|20 | ···················· | ···················· | ···················· | ···················· | ···················· | ···················· |
| O | 5969 hCoV-19/England/MILK-A7B17C/2020\|EPI_ISL_607552\|20 | ···················· | ···················· | ···················· | ···················· | ···················· | ···················· |
| O | 5970 hCoV-19/England/MILK-A7C8D0/2020\|EPI_ISL_607553\|20 | ···················· | ·A·················· | ···················· | ···················· | ···················· | ···A················ |
| O | 5971 hCoV-19/England/MILK-A7BC4E/2020\|EPI_ISL_607554\|20 | ···················· | ···················· | ···················· | ···················· | ······T············· | ···············A···· |
| O | 5972 hCoV-19/England/MILK-A7B570/2020\|EPI_ISL_607555\|20 | ···················· | ···················· | ···················· | ···················· | ···················· | ···················· |
| O | 5973 hCoV-19/England/MILK-A7BDC3/2020\|EPI_ISL_607556\|20 | ···················· | ···················· | ···················· | ···················· | ···················· | ···················· |
| O | 5974 hCoV-19/England/MILK-A7BADB/2020\|EPI_ISL_607557\|20 | ···················· | ···················· | ···················· | ···················· | ···················· | ···················· |
| O | 5975 hCoV-19/England/MILK-A7B428/2020\|EPI_ISL_607558\|20 | ···················· | ·A·················· | ···················· | ···················· | ···················· | ···A················ |
| O | 5976 hCoV-19/England/MILK-A7C937/2020\|EPI_ISL_607559\|20 | ···················· | ···················· | ···················· | ···················· | ···················· | ···················· |
| O | 5977 hCoV-19/England/MILK-A7BC3F/2020\|EPI_ISL_607560\|20 | ···················· | ···················· | ···················· | ···················· | ···················· | ···················· |
| O | 5978 hCoV-19/England/MILK-A7BB8D/2020\|EPI_ISL_607561\|20 | ···················· | ···················· | ···················· | ···················· | ···················· | ···················· |
| O | 5979 hCoV-19/England/MILK-A7B23D/2020\|EPI_ISL_607563\|20 | ···················· | ···················· | ···················· | ···················· | ···················· | ···················· |
| O | 5980 hCoV-19/England/MILK-A7BC11/2020\|EPI_ISL_607564\|20 | ···················· | ···················· | ···················· | ···················· | ···················· | ···················· |
| O | 5981 hCoV-19/England/MILK-A7BFEB/2020\|EPI_ISL_607565\|20 | ···················· | ·A·················· | ···················· | ···················· | ···················· | ···A················ |
| O | 5982 hCoV-19/England/MILK-A7AFBF/2020\|EPI_ISL_607567\|20 | ···················· | ···················· | ···················· | ···················· | ···················· | ···················· |
| O | 5983 hCoV-19/England/MILK-A7BC7B/2020\|EPI_ISL_607568\|20 | ···················· | ···················· | ···················· | ···················· | ···················· | ···················· |
| O | 5984 hCoV-19/England/MILK-A7C1B7/2020\|EPI_ISL_607569\|20 | ···················· | ···················· | ···················· | ···················· | ···················· | ···················· |
| O | 5985 hCoV-19/England/MILK-A7B543/2020\|EPI_ISL_607570\|20 | ···················· | ···················· | ···················· | ···················· | ···················· | ···················· |
| O | 5986 hCoV-19/England/MILK-A7C964/2020\|EPI_ISL_607571\|20 | ···················· | ·A·················· | ···················· | ···················· | ···················· | ···A················ |
| O | 5987 hCoV-19/England/MILK-A7B2F1/2020\|EPI_ISL_607573\|20 | ···················· | ···················· | ···················· | ···················· | ···················· | ···················· |
| O | 5988 hCoV-19/England/MILK-A7B121/2020\|EPI_ISL_607574\|20 | ···················· | ···················· | ···················· | ···················· | ···················· | ···················· |
| O | 5989 hCoV-19/England/MILK-A7C8FE/2020\|EPI_ISL_607576\|20 | ···················· | ···················· | ···················· | ···················· | ···················· | ···················· |
| O | 5990 hCoV-19/England/MILK-A7BCE4/2020\|EPI_ISL_607577\|20 | ···················· | ·A·················· | ···················· | ···················· | ···················· | ···················· |
| O | 5991 hCoV-19/England/MILK-A7B6B9/2020\|EPI_ISL_607578\|20 | ···················· | ···················· | ···················· | ···················· | ···················· | ···················· |
| O | 5992 hCoV-19/England/MILK-A7B0AC/2020\|EPI_ISL_607580\|20 | ···················· | ···················· | ···················· | ···················· | ···················· | ···················· |
| O | 5993 hCoV-19/England/MILK-A7C445/2020\|EPI_ISL_607581\|20 | ···················· | ···················· | ···················· | ···················· | ···················· | ···················· |
| O | 5994 hCoV-19/England/MILK-A7B2D3/2020\|EPI_ISL_607582\|20 | ···················· | ···················· | ···················· | ···················· | ···················· | ···················· |
| O | 5995 hCoV-19/England/MILK-A7C797/2020\|EPI_ISL_607584\|20 | ···················· | ·A·················· | ···················· | ···················· | ···················· | ···A················ |
| O | 5996 hCoV-19/England/MILK-A7AF55/2020\|EPI_ISL_607585\|20 | ···················· | ···················· | ···················· | ···················· | ···················· | ···················· |
| O | 5997 hCoV-19/England/MILK-A7C15D/2020\|EPI_ISL_607588\|20 | ···················· | ···················· | ···················· | ···················· | ···················· | ···················· |
| O | 5998 hCoV-19/England/MILK-A7B30D/2020\|EPI_ISL_607589\|20 | ···················· | ···················· | ···················· | ···················· | ···················· | ···················· |
| O | 5999 hCoV-19/England/MILK-A7B8B3/2020\|EPI_ISL_607590\|20 | ···················· | ···················· | ···················· | ···················· | ···················· | ···················· |
| O | 6000 hCoV-19/England/MILK-A7B446/2020\|EPI_ISL_607591\|20 | ···················· | ···················· | ···················· | ···················· | ···················· | ···················· |
| O | 6001 hCoV-19/England/MILK-A7BFBE/2020\|EPI_ISL_607592\|20 | ···················· | ···················· | ···················· | ···················· | ···················· | ···················· |
| O | 6002 hCoV-19/England/MILK-A7B09D/2020\|EPI_ISL_607593\|20 | ···················· | ·A·················· | ···················· | ···················· | ···················· | ···A················ |
| O | 6003 hCoV-19/England/MILK-A7C885/2020\|EPI_ISL_607594\|20 | ···················· | ···················· | ···················· | ···················· | ···················· | ···················· |
| O | 6004 hCoV-19/England/MILK-A7BAAE/2020\|EPI_ISL_607595\|20 | ···················· | ···················· | ···················· | ···················· | ···················· | ···················· |
| O | 6005 hCoV-19/England/MILK-A7BBBA/2020\|EPI_ISL_607596\|20 | ···················· | ···················· | ···················· | ···················· | ···················· | ···················· |

| | | | | | | | | |
|---|---|---|---|---|---|---|---|---|
| O | 6006 | hCoV-19/England/MILK-A7B516/2020\|EPI_ISL_607597\|20 | ···················· | ···················· | ···················· | ···················· | ···················· | ···················· |
| O | 6007 | hCoV-19/England/MILK-A7CBE6/2020\|EPI_ISL_607598\|20 | ···················· | ···················· | ···················· | ···················· | ···················· | ···················· |
| O | 6008 | hCoV-19/England/MILK-A7B0BB/2020\|EPI_ISL_607599\|20 | ···················· | ···················· | ···················· | ···················· | ···················· | ···················· |
| O | 6009 | hCoV-19/England/MILK-A7B3FE/2020\|EPI_ISL_607600\|20 | ···················· | ···················· | ···················· | ···················· | ···················· | ···················· |
| O | 6010 | hCoV-19/England/MILK-A7B9CF/2020\|EPI_ISL_607601\|20 | ···················· | ···················· | ···················· | ···················· | ···················· | ···················· |
| O | 6011 | hCoV-19/England/MILK-A7CACB/2020\|EPI_ISL_607602\|20 | ···················· | ···················· | ···················· | ···················· | ···················· | ···················· |
| O | 6012 | hCoV-19/England/MILK-A7C506/2020\|EPI_ISL_607603\|20 | ···················· | ···················· | ···················· | ···················· | ···················· | ···················· |
| O | 6013 | hCoV-19/England/MILK-A7B604/2020\|EPI_ISL_607604\|20 | ···················· | ···················· | ···················· | ···················· | ······T············· | ···················· |
| O | 6014 | hCoV-19/England/MILK-A7C849/2020\|EPI_ISL_607605\|20 | ···················· | ···················· | ···················· | ···················· | ···················· | ···················· |
| O | 6015 | hCoV-19/England/MILK-A7B67D/2020\|EPI_ISL_607606\|20 | ···················· | ···················· | ···················· | ···················· | ···················· | ···················· |
| O | 6016 | hCoV-19/England/MILK-A7C982/2020\|EPI_ISL_607607\|20 | ···················· | ···················· | ···················· | ···················· | ···················· | ···················· |
| O | 6017 | hCoV-19/England/MILK-A7B929/2020\|EPI_ISL_607608\|20 | ···················· | ···················· | ···················· | ···················· | ···················· | ···················· |
| O | 6018 | hCoV-19/England/MILK-A7BEB1/2020\|EPI_ISL_607609\|20 | ···················· | ···················· | ···················· | ···················· | ···················· | ···················· |
| O | 6019 | hCoV-19/England/MILK-A7B042/2020\|EPI_ISL_607610\|20 | ···················· | ···················· | ···················· | ···················· | ···················· | ···················· |
| O | 6020 | hCoV-19/England/MILK-A7B8C2/2020\|EPI_ISL_607611\|20 | ···················· | ···················· | ···················· | ···················· | ···················· | ···················· |
| O | 6021 | hCoV-19/England/MILK-A7B877/2020\|EPI_ISL_607613\|20 | ···················· | ···················· | ···················· | ···················· | ···················· | ···················· |
| O | 6022 | hCoV-19/England/MILK-A7C8A3/2020\|EPI_ISL_607615\|20 | ···················· | ···················· | ···················· | ···················· | ······T············· | ···················· |
| O | 6023 | hCoV-19/England/MILK-A7BE57/2020\|EPI_ISL_607616\|20 | ···················· | ·A·················· | ···················· | ···················· | ···················· | ··A················· |
| O | 6024 | hCoV-19/England/MILK-A7BC99/2020\|EPI_ISL_607617\|20 | ···················· | ···················· | ···················· | ···················· | ···················· | ···················· |
| O | 6025 | hCoV-19/England/MILK-A7B4CE/2020\|EPI_ISL_607618\|20 | ···················· | ···················· | ···················· | ···················· | ···················· | ···················· |
| O | 6026 | hCoV-19/England/MILK-A7C67C/2020\|EPI_ISL_607619\|20 | ···················· | ···················· | ···················· | ···················· | ···················· | ···················· |
| O | 6027 | hCoV-19/England/MILK-A7BB14/2020\|EPI_ISL_607620\|20 | ···················· | ·A·················· | ···················· | ···················· | ···················· | ··A················· |
| O | 6028 | hCoV-19/England/MILK-A7B394/2020\|EPI_ISL_607621\|20 | ···················· | ···················· | ···················· | ···················· | ···················· | ···················· |
| O | 6029 | hCoV-19/England/MILK-A7C9CE/2020\|EPI_ISL_607622\|20 | ···················· | ···················· | ···················· | ···················· | ···················· | ···················· |
| O | 6030 | hCoV-19/England/ALDP-A6F542/2020\|EPI_ISL_607623\|20 | ···················· | ···················· | ···················· | ···················· | ···················· | ··A················· |
| O | 6031 | hCoV-19/England/ALDP-A6DB33/2020\|EPI_ISL_607624\|20 | ···················· | ···················· | ···················· | ···················· | ···················· | ···················· |
| O | 6032 | hCoV-19/England/ALDP-A6E789/2020\|EPI_ISL_607625\|20 | ···················· | ···················· | ···················· | ···················· | ···················· | ···················· |
| O | 6033 | hCoV-19/England/ALDP-A6F4CD/2020\|EPI_ISL_607626\|20 | ···················· | ···················· | ···················· | ···················· | ···················· | ···················· |
| O | 6034 | hCoV-19/England/ALDP-A6DBE8/2020\|EPI_ISL_607628\|20 | ···················· | ···················· | ···················· | ···················· | ···················· | ···················· |
| O | 6035 | hCoV-19/England/ALDP-A700B6/2020\|EPI_ISL_607629\|20 | ···················· | ···················· | ···················· | ···················· | ···················· | ···················· |
| O | 6036 | hCoV-19/England/ALDP-A70502/2020\|EPI_ISL_607651\|20 | ···················· | ···················· | ···················· | ···················· | ···················· | ···················· |
| O | 6037 | hCoV-19/England/ALDP-A6EF72/2020\|EPI_ISL_607661\|20 | ···················· | ···················· | ···················· | ···················· | ···················· | ···················· |
| O | 6038 | hCoV-19/England/ALDP-A6E0CA/2020\|EPI_ISL_607662\|20 | ···················· | ···················· | ···················· | ···················· | ···················· | ···················· |
| O | 6039 | hCoV-19/England/ALDP-A6E060/2020\|EPI_ISL_607663\|20 | ···················· | ···················· | ···················· | ···················· | ···················· | ···················· |
| O | 6040 | hCoV-19/England/ALDP-A6DA36/2020\|EPI_ISL_607664\|20 | ···················· | ···················· | ···················· | ···················· | ···················· | ···················· |
| O | 6041 | hCoV-19/England/ALDP-A6F13F/2020\|EPI_ISL_607665\|20 | ···················· | ···················· | ···················· | ···················· | ···················· | ···················· |
| O | 6042 | hCoV-19/England/ALDP-A6F67C/2020\|EPI_ISL_607666\|20 | ···················· | ···················· | ···················· | ···················· | ···················· | ···················· |
| O | 6043 | hCoV-19/England/ALDP-A70308/2020\|EPI_ISL_607667\|20 | ···················· | ···················· | ···················· | ···················· | ···················· | ···················· |
| O | 6044 | hCoV-19/England/ALDP-A6E3FE/2020\|EPI_ISL_607669\|20 | ···················· | ···················· | ···················· | ···················· | ···················· | ···················· |
| O | 6045 | hCoV-19/England/ALDP-A6919F/2020\|EPI_ISL_608880\|20 | ···················· | ···················· | ···················· | ···················· | ···················· | ···················· |
| O | 6046 | hCoV-19/England/MILK-A6C080/2020\|EPI_ISL_608882\|20 | ···················· | ···················· | ···················· | ···················· | ···················· | ···················· |
| O | 6047 | hCoV-19/England/ALDP-A68B0B/2020\|EPI_ISL_608883\|20 | ···················· | ···················· | ···················· | ···················· | ···················· | ···················· |
| O | 6048 | hCoV-19/England/ALDP-A6828E/2020\|EPI_ISL_608884\|20 | ···················· | ···················· | ···················· | ···················· | ···················· | ···················· |
| O | 6049 | hCoV-19/England/ALDP-A69478/2020\|EPI_ISL_608885\|20 | ···················· | ···················· | ···················· | ···················· | ···················· | ···················· |
| O | 6050 | hCoV-19/England/ALDP-A6926F/2020\|EPI_ISL_608886\|20 | ···················· | ···················· | ···················· | ···················· | ···················· | ···················· |
| O | 6051 | hCoV-19/England/ALDP-A682BB/2020\|EPI_ISL_608888\|20 | ···················· | ···················· | ···················· | ···················· | ···················· | ···················· |
| O | 6052 | hCoV-19/England/ALDP-A6800C/2020\|EPI_ISL_608889\|20 | ···················· | ···················· | ···················· | ···················· | ···················· | ···················· |
| O | 6053 | hCoV-19/England/ALDP-A69144/2020\|EPI_ISL_608890\|20 | ···················· | ···················· | ···················· | ···················· | ···················· | ···················· |
| O | 6054 | hCoV-19/England/ALDP-A67D51/2020\|EPI_ISL_608891\|20 | ···················· | ···················· | ···················· | ···················· | ···················· | ···················· |
| O | 6055 | hCoV-19/England/CAMC-A64686/2020\|EPI_ISL_608892\|20 | ···················· | ···················· | ···················· | ···················· | ···················· | ···················· |
| O | 6056 | hCoV-19/England/MILK-A6BF84/2020\|EPI_ISL_608893\|20 | ···················· | ···················· | ···················· | ···················· | ···················· | ···················· |
| O | 6057 | hCoV-19/England/ALDP-A6945A/2020\|EPI_ISL_608895\|20 | ···················· | ···················· | ···················· | ···················· | ···················· | ···················· |
| O | 6058 | hCoV-19/England/ALDP-A68136/2020\|EPI_ISL_608897\|20 | ···················· | ···················· | ···················· | ···················· | ···················· | ···················· |
| O | 6059 | hCoV-19/England/ALDP-A68AC2/2020\|EPI_ISL_608898\|20 | ···················· | ···················· | ···················· | ···················· | ···················· | ···················· |
| O | 6060 | hCoV-19/England/ALDP-A69399/2020\|EPI_ISL_608899\|20 | ···················· | ···················· | ···················· | ···················· | ···················· | ···················· |
| O | 6061 | hCoV-19/England/CAMC-A64C57/2020\|EPI_ISL_608918\|20 | ···················· | ···················· | ···················· | ···················· | ···················· | ···················· |
| O | 6062 | hCoV-19/England/ALDP-A68FF0/2020\|EPI_ISL_608919\|20 | ···················· | ···················· | ···················· | ···················· | ···················· | ···················· |
| O | 6063 | hCoV-19/England/ALDP-A67D15/2020\|EPI_ISL_608920\|20 | ···················· | ···················· | ···················· | ···················· | ···················· | ···················· |
| O | 6064 | hCoV-19/England/ALDP-A69250/2020\|EPI_ISL_608921\|20 | ···················· | ···················· | ···················· | ···················· | ···················· | ···················· |
| O | 6065 | hCoV-19/England/ALDP-A69223/2020\|EPI_ISL_608922\|20 | ···················· | ···················· | ···················· | ···················· | ···················· | ···················· |
| O | 6066 | hCoV-19/England/CAMC-A645C5/2020\|EPI_ISL_608923\|20 | ···················· | ···················· | ···················· | ···················· | ···················· | ···················· |
| O | 6067 | hCoV-19/England/CAMC-A64AB7/2020\|EPI_ISL_608924\|20 | ···················· | ···················· | ···················· | ···················· | ···················· | ···················· |
| O | 6068 | hCoV-19/England/ALDP-A691AE/2020\|EPI_ISL_608925\|20 | ···················· | ···················· | ···················· | ···················· | ···················· | ···················· |
| O | 6069 | hCoV-19/England/ALDP-A68066/2020\|EPI_ISL_608926\|20 | ···················· | ···················· | ···················· | ···················· | ···················· | ···················· |
| O | 6070 | hCoV-19/England/CAMC-A6555B/2020\|EPI_ISL_608927\|20 | ···················· | ···················· | ···················· | ···················· | ···················· | ···················· |
| O | 6071 | hCoV-19/England/ALDP-A69496/2020\|EPI_ISL_608928\|20 | ···················· | ···················· | ···················· | ···················· | ···················· | ···················· |
| O | 6072 | hCoV-19/England/ALDP-A695A2/2020\|EPI_ISL_608929\|20 | ···················· | ···················· | ···················· | ···················· | ···················· | ···················· |
| O | 6073 | hCoV-19/Wales/ALDP-A68A0E/2020\|EPI_ISL_608930\|2020 | ···················· | ···················· | ···················· | ···················· | ···················· | ···················· |
| O | 6074 | hCoV-19/England/ALDP-A69135/2020\|EPI_ISL_608931\|20 | ···················· | ···················· | ···················· | ···················· | ···················· | ···················· |
| O | 6075 | hCoV-19/England/ALDP-A69092/2020\|EPI_ISL_608932\|20 | ···················· | ···················· | ···················· | ···················· | ···················· | ···················· |
| O | 6076 | hCoV-19/England/ALDP-A68A95/2020\|EPI_ISL_608933\|20 | ···················· | ···················· | ···················· | ···················· | ···················· | ···················· |
| O | 6077 | hCoV-19/England/ALDP-A69311/2020\|EPI_ISL_608934\|20 | ···················· | ···················· | ···················· | ···················· | ···················· | ···················· |
| O | 6078 | hCoV-19/England/CAMC-A6457A/2020\|EPI_ISL_608935\|20 | ···················· | ···················· | ···················· | ···················· | ···················· | ···················· |
| O | 6079 | hCoV-19/England/ALDP-A69557/2020\|EPI_ISL_608938\|20 | ···················· | ···················· | ···················· | ···················· | ···················· | ···················· |
| O | 6080 | hCoV-19/England/CAMC-A64CA2/2020\|EPI_ISL_608939\|20 | ···················· | ···················· | ···················· | ···················· | ···················· | ···················· |
| O | 6081 | hCoV-19/England/MILK-A6BDF3/2020\|EPI_ISL_608940\|20 | ···················· | ···················· | ···················· | ···················· | ···················· | ···················· |
| O | 6082 | hCoV-19/England/ALDP-A69029/2020\|EPI_ISL_608941\|20 | ···················· | ···················· | ···················· | ···················· | ···················· | ···················· |
| O | 6083 | hCoV-19/England/CAMC-A64932/2020\|EPI_ISL_608943\|20 | ···················· | ···················· | ···················· | ···················· | ···················· | ···················· |
| O | 6084 | hCoV-19/England/ALDP-A69654/2020\|EPI_ISL_608944\|20 | ···················· | ···················· | ···················· | ···················· | ···················· | ···················· |
| O | 6085 | hCoV-19/England/ALDP-A682E8/2020\|EPI_ISL_608945\|20 | ···················· | ···················· | ···················· | ···················· | ···················· | ···················· |

| | | | | | | | | |
|---|---|---|---|---|---|---|---|---|
| O | 6086 | hCoV-19/England/ALDP-A69241/2020\|EPI_ISL_608946\|20 | ···················· | ···················· | ····················· | ···················· | ···················· | ···················· |
| O | 6087 | hCoV-19/England/CAMC-A645F2/2020\|EPI_ISL_608947\|20 | ···················· | ···················· | ····················· | ···················· | ···················· | ···················· |
| O | 6088 | hCoV-19/England/ALDP-A691EA/2020\|EPI_ISL_608948\|20 | ···················· | ···················· | ····················· | ···················· | ···················· | ···················· |
| O | 6089 | hCoV-19/England/ALDP-A6935D/2020\|EPI_ISL_608950\|20 | ···················· | ···················· | ····················· | ···················· | ···················· | ···················· |
| O | 6090 | hCoV-19/England/MILK-A6BF0C/2020\|EPI_ISL_608951\|20 | ···················· | ···················· | ····················· | ···················· | ···················· | ···················· |
| O | 6091 | hCoV-19/England/ALDP-A6829D/2020\|EPI_ISL_608952\|20 | ···················· | ···················· | ····················· | ···················· | ···················· | ···················· |
| O | 6092 | hCoV-19/England/MILK-A6BFEE/2020\|EPI_ISL_608954\|20 | ···················· | ·A·················· | ····················· | ···················· | ···················· | ··A················· |
| O | 6093 | hCoV-19/England/ALDP-A692F6/2020\|EPI_ISL_608956\|20 | ···················· | ···················· | ····················· | ···················· | ···················· | ···················· |
| O | 6094 | hCoV-19/England/ALDP-A68A3B/2020\|EPI_ISL_608957\|20 | ···················· | ···················· | ····················· | ···················· | ···················· | ···················· |
| O | 6095 | hCoV-19/England/CAMC-A645D4/2020\|EPI_ISL_608958\|20 | ···················· | ···················· | ····················· | ···················· | ···················· | ···················· |
| O | 6096 | hCoV-19/England/CAMC-A64A6C/2020\|EPI_ISL_608959\|20 | ···················· | ···················· | ····················· | ···················· | ···················· | ···················· |
| O | 6097 | hCoV-19/England/ALDP-A6941E/2020\|EPI_ISL_608980\|20 | ···················· | ···················· | ····················· | ···················· | ···················· | ···················· |
| O | 6098 | hCoV-19/England/CAMC-A645B6/2020\|EPI_ISL_608982\|20 | ···················· | ···················· | ····················· | ···················· | ···················· | ···················· |
| O | 6099 | hCoV-19/England/CAMC-A64950/2020\|EPI_ISL_608990\|20 | ···················· | ·A·······A·········· | ····················· | ···················· | ···················· | ···················· |
| O | 6100 | hCoV-19/England/ALDP-A68251/2020\|EPI_ISL_608991\|20 | ···················· | ···················· | ····················· | ···················· | ···················· | ···················· |
| O | 6101 | hCoV-19/England/ALDP-A68BCF/2020\|EPI_ISL_608992\|20 | ···················· | ···················· | ····················· | ···················· | ···················· | ···················· |
| O | 6102 | hCoV-19/England/ALDP-A68075/2020\|EPI_ISL_608994\|20 | ···················· | ···················· | ····················· | ···················· | ···················· | ···················· |
| O | 6103 | hCoV-19/England/CAMC-A64A5D/2020\|EPI_ISL_608996\|20 | ···················· | ···················· | ····················· | ···················· | ···················· | ···················· |
| O | 6104 | hCoV-19/England/CAMC-A6489F/2020\|EPI_ISL_608997\|20 | ···················· | ···················· | ····················· | ···················· | ···················· | ···················· |
| O | 6105 | hCoV-19/England/ALDP-A695FD/2020\|EPI_ISL_608998\|20 | ···················· | ···················· | ····················· | ···················· | ···················· | ···················· |
| O | 6106 | hCoV-19/England/ALDP-A69083/2020\|EPI_ISL_608999\|20 | ···················· | ···················· | ····················· | ···················· | ···················· | ···················· |
| O | 6107 | hCoV-19/England/CAMB-1B6D10/2020\|EPI_ISL_612186\|20 | ···················· | ···················· | ····················· | ···················· | ······T············· | ···················· |
| O | 6108 | hCoV-19/Scotland/EDB8376/2020\|EPI_ISL_612187\|2020- | ···················· | ···················· | ····················· | ···················· | ···················· | ···················· |
| O | 6110 | hCoV-19/Scotland/EDB8430/2020\|EPI_ISL_612189\|2020- | ···················· | ···················· | ····················· | ···················· | ···················· | ···················· |
| O | 6111 | hCoV-19/England/LIVE-1D2A62/2020\|EPI_ISL_612190\|20 | ···················· | ···················· | ····················· | ···················· | ···················· | ···················· |
| O | 6112 | hCoV-19/England/LIVE-1D2E66/2020\|EPI_ISL_612191\|20 | ···················· | ···················· | ····················· | ···················· | ···················· | ···················· |
| O | 6113 | hCoV-19/England/NOTT-117445/2020\|EPI_ISL_612192\|20 | ···················· | ···················· | ····················· | ···················· | ···················· | ···················· |
| O | 6114 | hCoV-19/Wales/PHWC-4809FA/2020\|EPI_ISL_612193\|2020 | ···················· | ···················· | ····················· | ···················· | ···················· | ···················· |
| O | 6116 | hCoV-19/Wales/PHWC-4822EE/2020\|EPI_ISL_612195\|2020 | ···················· | ···················· | ····················· | ···················· | ···················· | ···················· |
| O | 6117 | hCoV-19/England/SHEF-C8B6D/2020\|EPI_ISL_612196\|202 | ···················· | ·A·················· | ····················· | ···················· | ···················· | ··A················· |
| O | 6118 | hCoV-19/England/MILK-A18997/2020\|EPI_ISL_612198\|20 | ···················· | ···················· | ····················· | ···················· | ···················· | ···················· |
| O | 6119 | hCoV-19/Wales/PHWC-482406/2020\|EPI_ISL_612199\|2020 | ···················· | ···················· | ····················· | ···················· | ···················· | ···················· |
| O | 6120 | hCoV-19/England/QEUH-A78045/2020\|EPI_ISL_613354\|20 | ···················· | ···················· | ····················· | ···················· | ···················· | ···················· |
| O | 6121 | hCoV-19/England/QEUH-A78054/2020\|EPI_ISL_613355\|20 | ···················· | ···················· | ····················· | ···················· | ···················· | ···················· |
| O | 6122 | hCoV-19/England/SHEF-C0FA5/2020\|EPI_ISL_613356\|202 | ···················· | ·A·················· | ····················· | ···················· | ···················· | ···················· |
| O | 6123 | hCoV-19/England/SHEF-C0FC3/2020\|EPI_ISL_613357\|202 | ···················· | ·A·················· | ····················· | ···················· | ···················· | ···················· |
| O | 6124 | hCoV-19/England/SHEF-C10A1/2020\|EPI_ISL_613358\|202 | ···················· | ···················· | ····················· | ···················· | ···················· | ···················· |
| O | 6125 | hCoV-19/England/SHEF-C1AD0/2020\|EPI_ISL_613359\|202 | ···················· | ···················· | ····················· | ···················· | ···················· | ···················· |
| O | 6126 | hCoV-19/England/SHEF-D192B/2020\|EPI_ISL_613400\|202 | ···················· | ···················· | ····················· | ···················· | ···················· | ···················· |
| O | 6127 | hCoV-19/England/SHEF-D1A0A/2020\|EPI_ISL_613401\|202 | ···················· | ···················· | ····················· | ···················· | ···················· | ···················· |
| O | 6128 | hCoV-19/England/SHEF-D1DB6/2020\|EPI_ISL_613402\|202 | ···················· | ···················· | ····················· | ···················· | ···················· | ···················· |
| O | 6129 | hCoV-19/England/SHEF-D1E1D/2020\|EPI_ISL_613403\|202 | ···················· | ···················· | ····················· | ···················· | ···················· | ···················· |
| O | 6130 | hCoV-19/England/SHEF-D226B/2020\|EPI_ISL_613404\|202 | ···················· | ···················· | ····················· | ···················· | ···················· | ···················· |
| O | 6131 | hCoV-19/England/SHEF-D27B7/2020\|EPI_ISL_613405\|202 | ···················· | ···················· | ····················· | ···················· | ···················· | ···················· |
| O | 6132 | hCoV-19/England/SHEF-D2966/2020\|EPI_ISL_613406\|202 | ···················· | ···················· | ····················· | ···················· | ···················· | ···················· |
| O | 6133 | hCoV-19/England/SHEF-D3060/2020\|EPI_ISL_613407\|202 | ···················· | ···················· | ····················· | ···················· | ···················· | ···················· |
| O | 6134 | hCoV-19/England/SHEF-D3419/2020\|EPI_ISL_613408\|202 | ···················· | ···················· | ····················· | ···················· | ···················· | ···················· |
| O | 6135 | hCoV-19/England/SHEF-D34FB/2020\|EPI_ISL_613409\|202 | ···················· | ···················· | ····················· | ···················· | ···················· | ···················· |
| O | 6136 | hCoV-19/Guadeloupe/IPG-7319/2020\|EPI_ISL_613419\|20 | ···················· | ···················· | ····················· | ···················· | ···················· | ···················· |
| O | 6137 | hCoV-19/Guadeloupe/IPG-7320/2020\|EPI_ISL_613420\|20 | ···················· | ···················· | ····················· | ···················· | ···················· | ···················· |
| O | 6138 | hCoV-19/Guadeloupe/IPG-7321/2020\|EPI_ISL_613421\|20 | ···················· | ···················· | ····················· | ···················· | ···················· | ···················· |
| O | 6139 | hCoV-19/Guadeloupe/IPG-7322/2020\|EPI_ISL_613422\|20 | ···················· | ···················· | ····················· | ···················· | ···················· | ···················· |
| O | 6140 | hCoV-19/Guadeloupe/IPG-7323/2020\|EPI_ISL_613423\|20 | ···················· | ···················· | ····················· | ···················· | ···················· | ···················· |
| O | 6141 | hCoV-19/Guadeloupe/IPG-7324/2020\|EPI_ISL_613424\|20 | ···················· | ···················· | ····················· | ···················· | ···················· | ···················· |
| O | 6142 | hCoV-19/Guadeloupe/IPG-7325/2020\|EPI_ISL_613425\|20 | ···················· | ···················· | ····················· | ···················· | ···················· | ···················· |
| O | 6143 | hCoV-19/Guadeloupe/IPG-7326/2020\|EPI_ISL_613426\|20 | ···················· | ···················· | ····················· | ···················· | ···················· | ···················· |
| O | 6144 | hCoV-19/Guadeloupe/IPG-7327/2020\|EPI_ISL_613427\|20 | ···················· | ···················· | ····················· | ···················· | ···················· | ···················· |
| O | 6145 | hCoV-19/England/ALDP-A87A27/2020\|EPI_ISL_606140\|20 | ···················· | ···················· | ····················· | ···················· | ···················· | ···················· |
| O | 6146 | hCoV-19/England/ALDP-A8778A/2020\|EPI_ISL_606141\|20 | ···················· | ···················· | ····················· | ···················· | ···················· | ···················· |
| O | 6147 | hCoV-19/England/ALDP-A8740B/2020\|EPI_ISL_606143\|20 | ···················· | ···················· | ····················· | ···················· | ···················· | ···················· |
| O | 6148 | hCoV-19/England/ALDP-A86F92/2020\|EPI_ISL_606144\|20 | ···················· | ···················· | ····················· | ···················· | ···················· | ···················· |
| O | 6149 | hCoV-19/England/ALDP-A87ACD/2020\|EPI_ISL_606145\|20 | ···················· | ···················· | ····················· | ···················· | ···················· | ···················· |
| O | 6150 | hCoV-19/England/ALDP-A87869/2020\|EPI_ISL_606146\|20 | ···················· | ···················· | ····················· | ···················· | ···················· | ···················· |
| O | 6151 | hCoV-19/England/ALDP-A860BD/2020\|EPI_ISL_606147\|20 | ···················· | ···················· | ····················· | ···················· | ···················· | ···················· |
| O | 6152 | hCoV-19/England/ALDP-A8792A/2020\|EPI_ISL_606148\|20 | ···················· | ···················· | ····················· | ···················· | ···················· | ···················· |
| O | 6153 | hCoV-19/England/ALDP-A87A36/2020\|EPI_ISL_606149\|20 | ···················· | ···················· | ····················· | ···················· | ···················· | ···················· |
| O | 6154 | hCoV-19/England/ALDP-A8715F/2020\|EPI_ISL_606150\|20 | ···················· | ···················· | ····················· | ···················· | ···················· | ···················· |
| O | 6155 | hCoV-19/England/ALDP-A87535/2020\|EPI_ISL_606151\|20 | ···················· | ···················· | ····················· | ···················· | ···················· | ···················· |
| O | 6156 | hCoV-19/England/ALDP-A87B7F/2020\|EPI_ISL_606152\|20 | ···················· | ···················· | ····················· | ···················· | ···················· | ···················· |
| O | 6157 | hCoV-19/England/ALDP-A873B3/2020\|EPI_ISL_606153\|20 | ···················· | ···················· | ····················· | ···················· | ···················· | ···················· |
| O | 6158 | hCoV-19/England/ALDP-A87CA9/2020\|EPI_ISL_606154\|20 | ···················· | ···················· | ····················· | ···················· | ···················· | ···················· |
| O | 6159 | hCoV-19/England/ALDP-A8731D/2020\|EPI_ISL_606156\|20 | ···················· | ···················· | ····················· | ···················· | ···················· | ···················· |
| O | 6160 | hCoV-19/England/ALDP-A87D1F/2020\|EPI_ISL_606157\|20 | ···················· | ···················· | ····················· | ···················· | ···················· | ···················· |
| O | 6161 | hCoV-19/England/ALDP-A87526/2020\|EPI_ISL_606158\|20 | ···················· | ···················· | ····················· | ···················· | ···················· | ···················· |
| O | 6162 | hCoV-19/England/ALDP-A87122/2020\|EPI_ISL_606159\|20 | ···················· | ···················· | ····················· | ···················· | ···················· | ···················· |
| O | 6163 | hCoV-19/England/ALDP-A86EE0/2020\|EPI_ISL_606160\|20 | ···················· | ···················· | ····················· | ···················· | ···················· | ···················· |
| O | 6164 | hCoV-19/England/ALDP-A876BA/2020\|EPI_ISL_606162\|20 | ···················· | ···················· | ····················· | ···················· | ···················· | ···················· |
| O | 6165 | hCoV-19/England/ALDP-A873FF/2020\|EPI_ISL_606200\|20 | ···················· | ···················· | ····················· | ···················· | ···················· | ···················· |
| O | 6166 | hCoV-19/England/ALDP-A870CB/2020\|EPI_ISL_606201\|20 | ···················· | ···················· | ····················· | ···················· | ···················· | ···················· |
| O | 6167 | hCoV-19/England/ALDP-A87E76/2020\|EPI_ISL_606202\|20 | ···················· | ···················· | ····················· | ···················· | ···················· | ···················· |

| | | | | | | | | |
|---|---|---|---|---|---|---|---|---|
| O | 6168 | hCoV-19/England/ALDP-A86F74/2020\|EPI_ISL_606203\|20 | ···················· | ···················· | ···················· | ···················· | ···················· | ···················· |
| O | 6169 | hCoV-19/England/ALDP-A86CB9/2020\|EPI_ISL_606204\|20 | ···················· | ···················· | ···················· | ···················· | ···················· | ···················· |
| O | 6170 | hCoV-19/England/ALDP-A8780F/2020\|EPI_ISL_606205\|20 | ···················· | ···················· | ···················· | ···················· | ···················· | ···················· |
| O | 6171 | hCoV-19/England/ALDP-A86615/2020\|EPI_ISL_606206\|20 | ···················· | ···················· | ···················· | ···················· | ···················· | ···················· |
| O | 6173 | hCoV-19/England/ALDP-A86F47/2020\|EPI_ISL_606209\|20 | ···················· | ···················· | ···················· | ···················· | ···················· | ···················· |
| O | 6174 | hCoV-19/England/ALDP-A87298/2020\|EPI_ISL_606210\|20 | ···················· | ···················· | ···················· | ···················· | ···················· | ···················· |
| O | 6175 | hCoV-19/England/ALDP-A879A2/2020\|EPI_ISL_606211\|20 | ···················· | ···················· | ···················· | ···················· | ···················· | ···················· |
| O | 6176 | hCoV-19/England/ALDP-A87A90/2020\|EPI_ISL_606212\|20 | ···················· | ···················· | ···················· | ···················· | ···················· | ···················· |
| O | 6177 | hCoV-19/England/ALDP-A8733B/2020\|EPI_ISL_606213\|20 | ···················· | ···················· | ···················· | ···················· | ···················· | ···················· |
| O | 6178 | hCoV-19/England/ALDP-A87F19/2020\|EPI_ISL_606214\|20 | ···················· | ···················· | ···················· | ···················· | ···················· | ···················· |
| O | 6179 | hCoV-19/England/ALDP-A876E7/2020\|EPI_ISL_606216\|20 | ···················· | ···················· | ···················· | ···················· | ···················· | ···················· |
| O | 6180 | hCoV-19/England/ALDP-A8625D/2020\|EPI_ISL_606217\|20 | ···················· | ···················· | ···················· | ···················· | ···················· | ···················· |
| O | 6181 | hCoV-19/England/ALDP-A86651/2020\|EPI_ISL_606218\|20 | ···················· | ···················· | ···················· | ···················· | ···················· | ···················· |
| O | 6182 | hCoV-19/England/ALDP-A87359/2020\|EPI_ISL_606219\|20 | ···················· | ···················· | ···················· | ···················· | ···················· | ···················· |
| O | 6183 | hCoV-19/England/MILK-A7BACC/2020\|EPI_ISL_607470\|20 | ···················· | ···················· | ···················· | ···················· | ···················· | ···················· |
| O | 6184 | hCoV-19/England/MILK-A7BD87/2020\|EPI_ISL_607471\|20 | ···················· | ·A·················· | ···················· | ···················· | ···················· | ···A················ |
| O | 6185 | hCoV-19/England/MILK-A7C82B/2020\|EPI_ISL_607472\|20 | ···················· | ···················· | ···················· | ···················· | ···················· | ···················· |
| O | 6186 | hCoV-19/England/MILK-A7CBD7/2020\|EPI_ISL_607473\|20 | ···················· | ···················· | ···················· | ···················· | ···················· | ···················· |
| O | 6187 | hCoV-19/England/MILK-A7B72F/2020\|EPI_ISL_607474\|20 | ···················· | ···················· | ···················· | ···················· | ···················· | ···················· |
| O | 6189 | hCoV-19/England/MILK-A7BA35/2020\|EPI_ISL_607477\|20 | ···················· | ···················· | ···················· | ···················· | ···················· | ···················· |
| O | 6190 | hCoV-19/England/MILK-A7C68B/2020\|EPI_ISL_607478\|20 | ···················· | ···················· | ···················· | ···················· | ······T············· | ···················· |
| O | 6191 | hCoV-19/England/MILK-A7BDF0/2020\|EPI_ISL_607479\|20 | ···················· | ···················· | ···················· | ···················· | ···················· | ···················· |
| O | 6192 | hCoV-19/England/MILK-A7B3D0/2020\|EPI_ISL_607480\|20 | ···················· | ···················· | ···················· | ···················· | ···················· | ···················· |
| O | 6193 | hCoV-19/England/MILK-A7BA26/2020\|EPI_ISL_607481\|20 | ···················· | ···················· | ···················· | ···················· | ···················· | ···················· |
| O | 6194 | hCoV-19/England/MILK-A7B07F/2020\|EPI_ISL_607482\|20 | ···················· | ···················· | ···················· | ···················· | ···················· | ···················· |
| O | 6196 | hCoV-19/England/MILK-A7B798/2020\|EPI_ISL_607484\|20 | ···················· | ·A·················· | ···················· | ···················· | ···················· | ···A················ |
| O | 6197 | hCoV-19/England/MILK-A7B895/2020\|EPI_ISL_607485\|20 | ···················· | ···················· | ···················· | ···················· | ···················· | ···················· |
| O | 6198 | hCoV-19/England/MILK-A7B2B5/2020\|EPI_ISL_607486\|20 | ···················· | ···················· | ···················· | ···················· | ···················· | ···················· |
| O | 6199 | hCoV-19/England/MILK-A7C17B/2020\|EPI_ISL_607487\|20 | ···················· | ···················· | ···················· | ···················· | ···················· | ···················· |
| O | 6200 | hCoV-19/England/MILK-A7C13F/2020\|EPI_ISL_607488\|20 | ···················· | ···················· | ···················· | ···················· | ···················· | ···················· |
| O | 6201 | hCoV-19/England/MILK-A7BBAB/2020\|EPI_ISL_607489\|20 | ···················· | ···················· | ···················· | ···················· | ···················· | ···················· |
| O | 6202 | hCoV-19/England/MILK-A7CC89/2020\|EPI_ISL_607490\|20 | ···················· | ···················· | ···················· | ···················· | ···················· | ···················· |
| O | 6203 | hCoV-19/England/MILK-A7C8EF/2020\|EPI_ISL_607492\|20 | ···················· | ···················· | ···················· | ···················· | ···················· | ···················· |
| O | 6204 | hCoV-19/England/MILK-A7BF54/2020\|EPI_ISL_607493\|20 | ···················· | ···················· | ···················· | ···················· | ···················· | ···················· |
| O | 6205 | hCoV-19/England/MILK-A7B5E9/2020\|EPI_ISL_607500\|20 | ···················· | ···················· | ···················· | ···················· | ···················· | ···················· |
| O | 6206 | hCoV-19/England/MILK-A7AF64/2020\|EPI_ISL_607501\|20 | ···················· | ···················· | ···················· | ···················· | ···················· | ···················· |
| O | 6207 | hCoV-19/England/MILK-A7AF91/2020\|EPI_ISL_607502\|20 | ···················· | ···················· | ···················· | ···················· | ···················· | ···················· |
| O | 6208 | hCoV-19/England/MILK-A7B5BC/2020\|EPI_ISL_607503\|20 | ···················· | ·A·················· | ···················· | ···················· | ···················· | ···················· |
| O | 6209 | hCoV-19/England/MILK-A7C6C7/2020\|EPI_ISL_607504\|20 | ···················· | ···················· | ···················· | ···················· | ···················· | ···················· |
| O | 6210 | hCoV-19/England/MILK-A7B24C/2020\|EPI_ISL_607507\|20 | ···················· | ···················· | ···················· | ···················· | ···················· | ···················· |
| O | 6211 | hCoV-19/England/MILK-A7CC6B/2020\|EPI_ISL_607508\|20 | ···················· | ···················· | ···················· | ···················· | ···················· | ···················· |
| O | 6212 | hCoV-19/England/MILK-A7B6E6/2020\|EPI_ISL_607509\|20 | ···················· | ···················· | ···················· | ···················· | ···················· | ···················· |
| O | 6213 | hCoV-19/England/MILK-A7C6A9/2020\|EPI_ISL_607510\|20 | ···················· | ···················· | ···················· | ···················· | ···················· | ···················· |
| O | 6214 | hCoV-19/England/MILK-A7BE66/2020\|EPI_ISL_607511\|20 | ···················· | ···················· | ···················· | ···················· | ···················· | ···················· |
| O | 6215 | hCoV-19/England/MILK-A7CC2F/2020\|EPI_ISL_607512\|20 | ···················· | ···················· | ···················· | ···················· | ···················· | ···················· |
| O | 6216 | hCoV-19/England/MILK-A7AF73/2020\|EPI_ISL_607513\|20 | ···················· | ···················· | ···················· | ···················· | ···················· | ···················· |
| O | 6217 | hCoV-19/England/MILK-A7B2E2/2020\|EPI_ISL_607514\|20 | ···················· | ···················· | ···················· | ···················· | ···················· | ···················· |
| O | 6218 | hCoV-19/England/MILK-A7B640/2020\|EPI_ISL_607515\|20 | ···················· | ···················· | ···················· | ···················· | ···················· | ···················· |
| O | 6219 | hCoV-19/England/MILK-A7B349/2020\|EPI_ISL_607517\|20 | ···················· | ···················· | ···················· | ···················· | ······T············· | ···················· |
| O | 6220 | hCoV-19/England/MILK-A7BE75/2020\|EPI_ISL_607518\|20 | ···················· | ···················· | ···················· | ···················· | ···················· | ···················· |
| O | 6221 | hCoV-19/England/MILK-A7C7B5/2020\|EPI_ISL_607520\|20 | ···················· | ···················· | ···················· | ···················· | ···················· | ···················· |
| O | 6222 | hCoV-19/England/MILK-A7B534/2020\|EPI_ISL_607521\|20 | ···················· | ···················· | ···················· | ···················· | ···················· | ···················· |
| O | 6223 | hCoV-19/England/MILK-A7C991/2020\|EPI_ISL_607522\|20 | ···················· | ···················· | ···················· | ···················· | ···················· | ···················· |
| O | 6224 | hCoV-19/England/MILK-A7CB04/2020\|EPI_ISL_607523\|20 | ···················· | ···················· | ···················· | ···················· | ···················· | ···················· |
| O | 6225 | hCoV-19/England/MILK-A7B84A/2020\|EPI_ISL_607524\|20 | ···················· | ···················· | ···················· | ···················· | ···················· | ···················· |
| O | 6226 | hCoV-19/England/MILK-A7C75B/2020\|EPI_ISL_607525\|20 | ···················· | ···················· | ···················· | ···················· | ···················· | ···················· |
| O | 6227 | hCoV-19/England/MILK-A7B288/2020\|EPI_ISL_607526\|20 | ···················· | ···················· | ···················· | ···················· | ···················· | ···················· |
| O | 6228 | hCoV-19/England/MILK-A7BFCD/2020\|EPI_ISL_607527\|20 | ···················· | ···················· | ···················· | ···················· | ···················· | ···················· |
| O | 6229 | hCoV-19/England/MILK-A7B4EC/2020\|EPI_ISL_607528\|20 | ···················· | ···················· | ···················· | ···················· | ···················· | ···················· |
| O | 6230 | hCoV-19/England/MILK-A7C6D6/2020\|EPI_ISL_607529\|20 | ···················· | ···················· | ···················· | ···················· | ···················· | ···················· |
| O | 6231 | hCoV-19/England/MILK-A7B91A/2020\|EPI_ISL_607530\|20 | ···················· | ···················· | ···················· | ···················· | ···················· | ···················· |
| O | 6232 | hCoV-19/England/MILK-A7C788/2020\|EPI_ISL_607531\|20 | ···················· | ···················· | ···················· | ···················· | ···················· | ···················· |
| O | 6233 | hCoV-19/England/MILK-A7B974/2020\|EPI_ISL_607532\|20 | ···················· | ·A·······A·········· | ···················· | ···················· | ···················· | ···················· |
| O | 6234 | hCoV-19/England/MILK-A7BDE1/2020\|EPI_ISL_607533\|20 | ···················· | ·A·················· | ···················· | ···················· | ···················· | ···A················ |
| O | 6235 | hCoV-19/England/MILK-A7B6F5/2020\|EPI_ISL_607534\|20 | ···················· | ···················· | ···················· | ···················· | ···················· | ···················· |
| O | 6236 | hCoV-19/England/MILK-A7AF82/2020\|EPI_ISL_607535\|20 | ···················· | ···················· | ···················· | ···················· | ···················· | ···················· |
| O | 6237 | hCoV-19/England/MILK-A7BD3C/2020\|EPI_ISL_607536\|20 | ···················· | ···················· | ···················· | ···················· | ···················· | ···················· |
| O | 6238 | hCoV-19/England/MILK-A7C919/2020\|EPI_ISL_607537\|20 | ···················· | ···················· | ···················· | ···················· | ······T············· | ···················· |
| O | 6239 | hCoV-19/England/MILK-A7BF72/2020\|EPI_ISL_607538\|20 | ···················· | ···················· | ···················· | ···················· | ···················· | ···················· |
| O | 6240 | hCoV-19/England/MILK-A7B1B8/2020\|EPI_ISL_607540\|20 | ···················· | ···················· | ···················· | ···················· | ···················· | ···················· |
| O | 6241 | hCoV-19/England/MILK-A7B15E/2020\|EPI_ISL_607541\|20 | ···················· | ···················· | ···················· | ···················· | ···················· | ···················· |
| O | 6242 | hCoV-19/England/MILK-A7B385/2020\|EPI_ISL_607542\|20 | ···················· | ···················· | ···················· | ···················· | ···················· | ···················· |
| O | 6243 | hCoV-19/England/MILK-A7B68C/2020\|EPI_ISL_607543\|20 | ···················· | ···················· | ···················· | ···················· | ······T············· | ···················· |
| O | 6244 | hCoV-19/England/MILK-A7B3C1/2020\|EPI_ISL_607544\|20 | ···················· | ···················· | ···················· | ···················· | ···················· | ···················· |
| O | 6245 | hCoV-19/England/MILK-A7C8B2/2020\|EPI_ISL_607545\|20 | ···················· | ···················· | ···················· | ···················· | ···················· | ···················· |
| O | 6246 | hCoV-19/England/MILK-A7B6D7/2020\|EPI_ISL_607546\|20 | ···················· | ···················· | ···················· | ···················· | ···················· | ···················· |
| O | 6247 | hCoV-19/England/MILK-A7B3B2/2020\|EPI_ISL_607548\|20 | ···················· | ···················· | ···················· | ···················· | ···················· | ···················· |
| O | 6248 | hCoV-19/England/MILK-A7C876/2020\|EPI_ISL_607549\|20 | ···················· | ····G··············· | ···················· | ···················· | ···················· | ···················· |
| O | 6249 | hCoV-19/England/MILK-A6B115/2020\|EPI_ISL_608800\|20 | ···················· | ···················· | ···················· | ···················· | ···················· | ···················· |
| O | 6250 | hCoV-19/England/MILK-A69BEC/2020\|EPI_ISL_608801\|20 | ···················· | ···················· | ···················· | ···················· | ···················· | ···················· |

O 6251 hCoV-19/England/MILK-A69CCB/2020|EPI_ISL_608802|20 ···················· ···················· ····················· ···················· ···················· ····················
O 6252 hCoV-19/England/MILK-A6B78C/2020|EPI_ISL_608803|20 ···················· ···················· ····················· ···················· ···················· ····················
O 6253 hCoV-19/England/MILK-A6B7E6/2020|EPI_ISL_608804|20 ···················· ···················· ····················· ···················· ···················· ····················
O 6254 hCoV-19/England/MILK-A6ABBE/2020|EPI_ISL_608805|20 ···················· ·A·················· ····················· ···················· ···················· ····················
O 6255 hCoV-19/England/MILK-A6B3F1/2020|EPI_ISL_608806|20 ···················· ···················· ····················· ···················· ···················· ····················
O 6256 hCoV-19/England/MILK-A6BB17/2020|EPI_ISL_608807|20 ···················· ···················· ····················· ···················· ···················· ····················
O 6257 hCoV-19/England/MILK-A6B564/2020|EPI_ISL_608808|20 ···················· ···················· ····················· ···················· ···················· ····················
O 6258 hCoV-19/England/MILK-A6BA47/2020|EPI_ISL_608810|20 ···················· ···················· ····················· ···················· ···················· ····················
O 6259 hCoV-19/England/MILK-A6B1E8/2020|EPI_ISL_608811|20 ···················· ···················· ····················· ···················· ···················· ····················
O 6260 hCoV-19/England/MILK-A6ADC7/2020|EPI_ISL_608812|20 ···················· ···················· ····················· ···················· ···················· ····················
O 6261 hCoV-19/England/MILK-A6AF58/2020|EPI_ISL_608813|20 ···················· ···················· ····················· ···················· ···················· ····················
O 6262 hCoV-19/England/MILK-A6ACBB/2020|EPI_ISL_608814|20 ···················· ···················· ····················· ···················· ···················· ····················
O 6263 hCoV-19/England/MILK-A6B528/2020|EPI_ISL_608815|20 ···················· ·A·················· ····················· ···················· ···················· ··A·················
O 6264 hCoV-19/Wales/MILK-A6B17F/2020|EPI_ISL_608816|2020 ···················· ···················· ····················· ···················· ···················· ····················
O 6265 hCoV-19/England/MILK-A6B24F/2020|EPI_ISL_608817|20 ···················· ···················· ····················· ···················· ···················· ····················
O 6266 hCoV-19/England/MILK-A69C43/2020|EPI_ISL_608818|20 ···················· ···················· ····················· ···················· ···················· ····················
O 6267 hCoV-19/England/MILK-A6B203/2020|EPI_ISL_608820|20 ···················· ···················· ····················· ···················· ···················· ····················
O 6268 hCoV-19/England/MILK-A6B467/2020|EPI_ISL_608821|20 ···················· ···················· ····················· ···················· ···················· ····················
O 6269 hCoV-19/England/MILK-A69F4A/2020|EPI_ISL_608822|20 ···················· ···················· ····················· ···················· ···················· ····················
O 6270 hCoV-19/England/MILK-A6B300/2020|EPI_ISL_608823|20 ···················· ···················· ····················· ···················· ···················· ····················
O 6271 hCoV-19/England/MILK-A69F3B/2020|EPI_ISL_608825|20 ···················· ···················· ····················· ···················· ···················· ····················
O 6272 hCoV-19/England/MILK-A6B160/2020|EPI_ISL_608826|20 ···················· ···················· ····················· ···················· ···················· ····················
O 6273 hCoV-19/England/MILK-A69D04/2020|EPI_ISL_608828|20 ···················· ···················· ····················· ···················· ···················· ····················
O 6274 hCoV-19/England/MILK-A6B5BF/2020|EPI_ISL_608830|20 ···················· ···················· ···········T········· ···················· ···················· ····················
O 6275 hCoV-19/England/MILK-A6B18E/2020|EPI_ISL_608831|20 ···················· ···················· ····················· ···················· ···················· ····················
O 6276 hCoV-19/England/MILK-A6B79B/2020|EPI_ISL_608833|20 ···················· ···················· ····················· ···················· ···················· ····················
O 6277 hCoV-19/England/MILK-A6B40D/2020|EPI_ISL_608837|20 ···················· ···················· ····················· ···················· ···················· ····················
O 6278 hCoV-19/England/MILK-A69C9E/2020|EPI_ISL_608838|20 ···················· ·A·················· ····················· ···················· ···················· ··A·················
O 6279 hCoV-19/England/MILK-A6B519/2020|EPI_ISL_608839|20 ···················· ···················· ····················· ···················· ···················· ····················
O 6280 hCoV-19/England/MILK-A69D5F/2020|EPI_ISL_608841|20 ···················· ···················· ····················· ···················· ···················· ····················
O 6281 hCoV-19/England/MILK-A69D8C/2020|EPI_ISL_608842|20 ···················· ···················· ····················· ···················· ···················· ····················
O 6282 hCoV-19/England/MILK-A6B1BB/2020|EPI_ISL_608844|20 ···················· ···················· ····················· ···················· ···················· ····················
O 6283 hCoV-19/England/CAMC-A648EA/2020|EPI_ISL_608845|20 ···················· ···················· ····················· ···················· ···················· ····················
O 6284 hCoV-19/England/MILK-A6C17E/2020|EPI_ISL_608846|20 ···················· ···················· ····················· ···················· ···················· ····················
O 6285 hCoV-19/England/ALDP-A69180/2020|EPI_ISL_608847|20 ···················· ···················· ····················· ···················· ···················· ····················
O 6286 hCoV-19/England/MILK-A6BC6F/2020|EPI_ISL_608848|20 ···················· ···················· ····················· ···················· ···················· ····················
O 6287 hCoV-19/England/ALDP-A6950C/2020|EPI_ISL_608849|20 ···················· ···················· ····················· ···················· ···················· ····················
O 6288 hCoV-19/England/MILK-A6C0EA/2020|EPI_ISL_608850|20 ···················· ···················· ····················· ···················· ···················· ····················
O 6289 hCoV-19/England/ALDP-A68A59/2020|EPI_ISL_608851|20 ···················· ···················· ····················· ···················· ···················· ····················
O 6290 hCoV-19/England/CAMC-A646B3/2020|EPI_ISL_608852|20 ···················· ···················· ····················· ···················· ···················· ····················
O 6291 hCoV-19/England/ALDP-A68303/2020|EPI_ISL_608853|20 ···················· ···················· ····················· ···················· ···················· ····················
O 6292 hCoV-19/Scotland/MILK-A6C132/2020|EPI_ISL_608854|2 ···················· ···················· ····················· ···················· ···················· ····················
O 6293 hCoV-19/England/ALDP-A692BA/2020|EPI_ISL_608856|20 ···················· ···················· ····················· ···················· ···················· ····················
O 6294 hCoV-19/England/MILK-A6BFC0/2020|EPI_ISL_608857|20 ···················· ···················· ····················· ···················· ···················· ····················
O 6295 hCoV-19/England/ALDP-A6835E/2020|EPI_ISL_608858|20 ···················· ···················· ····················· ···················· ···················· ····················
O 6296 hCoV-19/England/MILK-A6C053/2020|EPI_ISL_608859|20 ···················· ···················· ····················· ···················· ···················· ····················
O 6297 hCoV-19/England/ALDP-A68A77/2020|EPI_ISL_608860|20 ···················· ···················· ····················· ···················· ···················· ····················
O 6298 hCoV-19/England/MILK-A6BD20/2020|EPI_ISL_608861|20 ···················· ···················· ····················· ···················· ···················· ····················
O 6299 hCoV-19/England/ALDP-A68A4A/2020|EPI_ISL_608862|20 ···················· ···················· ····················· ···················· ···················· ····················
O 6300 hCoV-19/England/ALDP-A68118/2020|EPI_ISL_608863|20 ···················· ···················· ····················· ···················· ···················· ····················
O 6301 hCoV-19/England/ALDP-A67D60/2020|EPI_ISL_608864|20 ···················· ···················· ····················· ···················· ···················· ····················
O 6302 hCoV-19/England/ALDP-A68AD1/2020|EPI_ISL_608865|20 ···················· ···················· ····················· ···················· ···················· ····················
O 6303 hCoV-19/England/ALDP-A692AB/2020|EPI_ISL_608866|20 ···················· ···················· ····················· ···················· ···················· ····················
O 6304 hCoV-19/England/CAMC-A64695/2020|EPI_ISL_608867|20 ···················· ···················· ····················· ···················· ···················· ····················
O 6305 hCoV-19/England/CAMC-A647CF/2020|EPI_ISL_608868|20 ···················· ···················· ····················· ···················· ···················· ····················
O 6306 hCoV-19/England/ALDP-A68AE0/2020|EPI_ISL_608869|20 ···················· ···················· ····················· ···················· ···················· ····················
O 6307 hCoV-19/England/MILK-A6BCF6/2020|EPI_ISL_608871|20 ···················· ···················· ····················· ···················· ···················· ····················
O 6308 hCoV-19/England/ALDP-A6942D/2020|EPI_ISL_608874|20 ···················· ···················· ····················· ···················· ···················· ····················
O 6309 hCoV-19/England/CAMC-A64783/2020|EPI_ISL_608875|20 ···················· ···················· ····················· ···················· ···················· ····················
O 6310 hCoV-19/England/CAMC-A6460E/2020|EPI_ISL_608876|20 ···················· ···················· ····················· ···················· ···················· ····················
O 6311 hCoV-19/England/ALDP-A682CA/2020|EPI_ISL_608877|20 ···················· ···················· ····················· ···················· ···················· ····················
O 6312 hCoV-19/England/ALDP-A68312/2020|EPI_ISL_608878|20 ···················· ···················· ····················· ···················· ···················· ····················
O 6313 hCoV-19/England/CAMC-A6496F/2020|EPI_ISL_608879|20 ···················· ···················· ····················· ···················· ···················· ····················
O 6314 hCoV-19/Scotland/QEUH-A75E7B/2020|EPI_ISL_613307|2 ···················· ···················· ····················· ···················· ···················· ····················
O 6315 hCoV-19/England/QEUH-A75E99/2020|EPI_ISL_613308|20 ···················· ···················· ····················· ···················· ···················· ····················
O 6316 hCoV-19/Scotland/QEUH-A75EA8/2020|EPI_ISL_613309|2 ···················· ···················· ····················· ···················· ···················· ····················
O 6317 hCoV-19/England/QEUH-A75EB7/2020|EPI_ISL_613310|20 ···················· ···················· ····················· ···················· ···················· ····················
O 6318 hCoV-19/Scotland/QEUH-A75EC6/2020|EPI_ISL_613311|2 ···················· ···················· ····················· ···················· ···················· ····················
O 6319 hCoV-19/England/QEUH-A75EE4/2020|EPI_ISL_613312|20 ···················· ···················· ····················· ···················· ···················· ····················
O 6320 hCoV-19/England/QEUH-A762C9/2020|EPI_ISL_613313|20 ···················· ···················· ····················· ···················· ···················· ··A·················
O 6321 hCoV-19/England/QEUH-A762D8/2020|EPI_ISL_613314|20 ···················· ·A·················· ····················· ···················· ···················· ····················
O 6322 hCoV-19/England/QEUH-A762F6/2020|EPI_ISL_613315|20 ···················· ···················· ····················· ···················· ···················· ····················
O 6323 hCoV-19/England/QEUH-A76302/2020|EPI_ISL_613316|20 ···················· ···················· ····················· ···················· ···················· ····················
O 6324 hCoV-19/England/QEUH-A76320/2020|EPI_ISL_613317|20 ···················· ···················· ····················· ···················· ···················· ····················
O 6325 hCoV-19/England/QEUH-A7634E/2020|EPI_ISL_613318|20 ···················· ···················· ····················· ···················· ······T············· ····················
O 6326 hCoV-19/England/QEUH-A7637B/2020|EPI_ISL_613319|20 ···················· ···················· ····················· ···················· ···················· ····················
O 6327 hCoV-19/England/QEUH-A763A8/2020|EPI_ISL_613320|20 ···················· ···················· ····················· ···················· ···················· ····················
O 6328 hCoV-19/England/QEUH-A763B7/2020|EPI_ISL_613321|20 ···················· ···················· ····················· ···················· ···················· ····················
O 6329 hCoV-19/England/QEUH-A763C6/2020|EPI_ISL_613322|20 ···················· ···················· ····················· ···················· ···················· ····················
O 6330 hCoV-19/England/QEUH-A763E4/2020|EPI_ISL_613323|20 ···················· ···················· ····················· ···················· ···················· ····················

| | | | | | | | | |
|---|---|---|---|---|---|---|---|---|
| O | 6331 | hCoV-19/England/QEUH-A763F3/2020\|EPI_ISL_613324\|20 | ···················· | ···················· | ···················· | ···················· | ···················· | ···················· |
| O | 6332 | hCoV-19/England/QEUH-A7642D/2020\|EPI_ISL_613325\|20 | ···················· | ···················· | ···················· | ···················· | ···················· | ···················· |
| O | 6333 | hCoV-19/England/QEUH-A7645A/2020\|EPI_ISL_613326\|20 | ···················· | ···················· | ···················· | ···················· | ···················· | ···················· |
| O | 6334 | hCoV-19/Scotland/QEUH-A766CD/2020\|EPI_ISL_613327\|2 | ···················· | ···················· | ···················· | ···················· | ···················· | ···················· |
| O | 6335 | hCoV-19/England/QEUH-A7679D/2020\|EPI_ISL_613328\|20 | ···················· | ···················· | ···················· | ···················· | ···················· | ···················· |
| O | 6336 | hCoV-19/England/QEUH-A767CA/2020\|EPI_ISL_613329\|20 | ···················· | ···················· | ···················· | ···················· | ···················· | ···················· |
| O | 6337 | hCoV-19/England/QEUH-A767D9/2020\|EPI_ISL_613330\|20 | ···················· | ···················· | ···················· | ···················· | ···················· | ···················· |
| O | 6338 | hCoV-19/England/QEUH-A767E8/2020\|EPI_ISL_613331\|20 | ···················· | ···················· | ···················· | ···················· | ···················· | ···················· |
| O | 6339 | hCoV-19/England/QEUH-A767F7/2020\|EPI_ISL_613332\|20 | ···················· | ···················· | ···················· | ···················· | ···················· | ···················· |
| O | 6340 | hCoV-19/England/QEUH-A76803/2020\|EPI_ISL_613333\|20 | ···················· | ···················· | ···················· | ···················· | ···················· | ···················· |
| O | 6341 | hCoV-19/England/QEUH-A76812/2020\|EPI_ISL_613334\|20 | ···················· | ···················· | ···················· | ···················· | ···················· | ···················· |
| O | 6342 | hCoV-19/England/QEUH-A76830/2020\|EPI_ISL_613335\|20 | ···················· | ···················· | ···················· | ···················· | ·····T·············· | ···················· |
| O | 6343 | hCoV-19/England/QEUH-A7694C/2020\|EPI_ISL_613336\|20 | ···················· | ···················· | ···················· | ···················· | ···················· | ···················· |
| O | 6344 | hCoV-19/England/QEUH-A7695B/2020\|EPI_ISL_613337\|20 | ···················· | ···················· | ···················· | ···················· | ···················· | ···················· |
| O | 6345 | hCoV-19/England/QEUH-A76979/2020\|EPI_ISL_613338\|20 | ···················· | ···················· | ···················· | ···················· | ···················· | ···················· |
| O | 6346 | hCoV-19/Scotland/QEUH-A76988/2020\|EPI_ISL_613339\|2 | ···················· | ···················· | ···················· | ···················· | ···················· | ···················· |
| O | 6347 | hCoV-19/Scotland/QEUH-A769A6/2020\|EPI_ISL_613340\|2 | ···················· | ···················· | ···················· | ···················· | ···················· | ···················· |
| O | 6348 | hCoV-19/Scotland/QEUH-A769D3/2020\|EPI_ISL_613341\|2 | ···················· | ···················· | ···················· | ···················· | ···················· | ···················· |
| O | 6349 | hCoV-19/Scotland/QEUH-A769E2/2020\|EPI_ISL_613342\|2 | ···················· | ···················· | ···················· | ···················· | ···················· | ···················· |
| O | 6350 | hCoV-19/Scotland/QEUH-A769F1/2020\|EPI_ISL_613343\|2 | ···················· | ···················· | ···················· | ···················· | ···················· | ···················· |
| O | 6351 | hCoV-19/Scotland/QEUH-A76A0D/2020\|EPI_ISL_613344\|2 | ···················· | ···················· | ···················· | ···················· | ···················· | ···················· |
| O | 6352 | hCoV-19/Scotland/QEUH-A76A2B/2020\|EPI_ISL_613345\|2 | ···················· | ···················· | ···················· | ···················· | ···················· | ···················· |
| O | 6353 | hCoV-19/Scotland/QEUH-A76A3A/2020\|EPI_ISL_613346\|2 | ···················· | ···················· | ···················· | ···················· | ···················· | ···················· |
| O | 6354 | hCoV-19/England/QEUH-A76C07/2020\|EPI_ISL_613347\|20 | ···················· | ···················· | ···················· | ···················· | ···················· | ···················· |
| O | 6355 | hCoV-19/England/QEUH-A77FB2/2020\|EPI_ISL_613348\|20 | ···················· | ···················· | ···················· | ···················· | ···················· | ···················· |
| O | 6356 | hCoV-19/England/QEUH-A77FC1/2020\|EPI_ISL_613349\|20 | ···················· | ···················· | ···················· | ···················· | ···················· | ···················· |
| O | 6357 | hCoV-19/England/QEUH-A77FEF/2020\|EPI_ISL_613350\|20 | ···················· | ···················· | ···················· | ···················· | ···················· | ···················· |
| O | 6358 | hCoV-19/England/QEUH-A78009/2020\|EPI_ISL_613351\|20 | ···················· | ···················· | ···················· | ···················· | ···················· | ···················· |
| O | 6359 | hCoV-19/England/QEUH-A78018/2020\|EPI_ISL_613352\|20 | ···················· | ···················· | ···················· | ···················· | ···················· | ···················· |
| O | 6360 | hCoV-19/England/QEUH-A78036/2020\|EPI_ISL_613353\|20 | ···················· | ···················· | ···················· | ···················· | ···················· | ···················· |
| O | 6361 | hCoV-19/England/SHEF-C2170/2020\|EPI_ISL_613360\|202 | ···················· | ·A·················· | ···················· | ···················· | ···················· | ···················· |
| O | 6362 | hCoV-19/England/SHEF-C340D/2020\|EPI_ISL_613361\|202 | ···················· | ···················· | ···················· | ···················· | ···················· | ···················· |
| O | 6363 | hCoV-19/England/SHEF-C3E3C/2020\|EPI_ISL_613362\|202 | ···················· | ···················· | ···················· | ···················· | ···················· | ···················· |
| O | 6364 | hCoV-19/England/SHEF-C418D/2020\|EPI_ISL_613363\|202 | ···················· | ···················· | ···················· | ···················· | ···················· | ···················· |
| O | 6365 | hCoV-19/England/SHEF-C4590/2020\|EPI_ISL_613364\|202 | ···················· | ···················· | ···················· | ···················· | ···················· | ···················· |
| O | 6366 | hCoV-19/England/SHEF-C5113/2020\|EPI_ISL_613365\|202 | ···················· | ···················· | ···················· | ···················· | ···················· | ···················· |
| O | 6367 | hCoV-19/England/SHEF-C534A/2020\|EPI_ISL_613366\|202 | ···················· | ···················· | ···················· | ···················· | ···················· | ···················· |
| O | 6368 | hCoV-19/England/SHEF-C5605/2020\|EPI_ISL_613367\|202 | ···················· | ···················· | ···················· | ···················· | ···················· | ···················· |
| O | 6369 | hCoV-19/England/SHEF-C5EEF/2020\|EPI_ISL_613368\|202 | ···················· | ···················· | ···················· | ···················· | ···················· | ···················· |
| O | 6370 | hCoV-19/England/SHEF-C624C/2020\|EPI_ISL_613369\|202 | ···················· | ···················· | ···················· | ···················· | ···················· | ···················· |
| O | 6371 | hCoV-19/England/SHEF-C6992/2020\|EPI_ISL_613370\|202 | ···················· | ·A·················· | ···················· | ···················· | ···················· | ···················· |
| O | 6372 | hCoV-19/England/SHEF-C6CA8/2020\|EPI_ISL_613371\|202 | ···················· | ···················· | ···················· | ···················· | ···················· | ···················· |
| O | 6373 | hCoV-19/England/SHEF-C74BE/2020\|EPI_ISL_613372\|202 | ···················· | ···················· | ···················· | ···················· | ···················· | ···················· |
| O | 6374 | hCoV-19/England/SHEF-C77E2/2020\|EPI_ISL_613373\|202 | ···················· | ·A·················· | ···················· | ···················· | ···················· | ···················· |
| O | 6375 | hCoV-19/England/SHEF-C865D/2020\|EPI_ISL_613374\|202 | ···················· | ···················· | ···················· | ···················· | ···················· | ···················· |
| O | 6376 | hCoV-19/England/SHEF-C8990/2020\|EPI_ISL_613375\|202 | ···················· | ···················· | ···················· | ···················· | ···················· | ···················· |
| O | 6377 | hCoV-19/England/SHEF-C9346/2020\|EPI_ISL_613376\|202 | ···················· | ···················· | ···················· | ···················· | ···················· | ···················· |
| O | 6378 | hCoV-19/England/SHEF-C9461/2020\|EPI_ISL_613377\|202 | ···················· | ···················· | ···················· | ···················· | ···················· | ···················· |
| O | 6379 | hCoV-19/England/SHEF-C95D7/2020\|EPI_ISL_613378\|202 | ···················· | ·A·················· | ···················· | ···················· | ···················· | ···················· |
| O | 6380 | hCoV-19/England/SHEF-C9917/2020\|EPI_ISL_613379\|202 | ···················· | ···················· | ···················· | ···················· | ···················· | ···················· |
| O | 6381 | hCoV-19/England/SHEF-C9A9C/2020\|EPI_ISL_613380\|202 | ···················· | ···················· | ···················· | ···················· | ···················· | ···················· |
| O | 6382 | hCoV-19/England/SHEF-CA239/2020\|EPI_ISL_613381\|202 | ···················· | ···················· | ···················· | ···················· | ···················· | ···················· |
| O | 6383 | hCoV-19/England/SHEF-CA336/2020\|EPI_ISL_613382\|202 | ···················· | ···················· | ···················· | ···················· | ···················· | ···················· |
| O | 6384 | hCoV-19/England/SHEF-CAB3E/2020\|EPI_ISL_613383\|202 | ···················· | ···················· | ···················· | ···················· | ···················· | ···················· |
| O | 6385 | hCoV-19/England/SHEF-CAFD8/2020\|EPI_ISL_613384\|202 | ···················· | ···················· | ···················· | ···················· | ···················· | ···················· |
| O | 6386 | hCoV-19/England/SHEF-CC2BF/2020\|EPI_ISL_613385\|202 | ···················· | ···················· | ···················· | ···················· | ···················· | ···················· |
| O | 6387 | hCoV-19/England/SHEF-CC677/2020\|EPI_ISL_613386\|202 | ···················· | ···················· | ···················· | ···················· | ···················· | ···················· |
| O | 6388 | hCoV-19/England/SHEF-CCDDC/2020\|EPI_ISL_613387\|202 | ···················· | ···················· | ···················· | ···················· | ···················· | ···················· |
| O | 6389 | hCoV-19/England/SHEF-CCE33/2020\|EPI_ISL_613388\|202 | ···················· | ···················· | ···················· | ···················· | ···················· | ···················· |
| O | 6390 | hCoV-19/England/SHEF-CD37F/2020\|EPI_ISL_613389\|202 | ···················· | ···················· | ···················· | ···················· | ···················· | ···················· |
| O | 6391 | hCoV-19/England/SHEF-CD658/2020\|EPI_ISL_613390\|202 | ···················· | ·A·················· | ···················· | ···················· | ···················· | ···················· |
| O | 6392 | hCoV-19/England/SHEF-CDAF2/2020\|EPI_ISL_613391\|202 | ···················· | ···················· | ···················· | ···················· | ···················· | ···················· |
| O | 6393 | hCoV-19/England/SHEF-CE1CF/2020\|EPI_ISL_613392\|202 | ···················· | ···················· | ···················· | ···················· | ···················· | ···················· |
| O | 6394 | hCoV-19/England/SHEF-CE727/2020\|EPI_ISL_613393\|202 | ···················· | ···················· | ···················· | ···················· | ···················· | ···················· |
| O | 6395 | hCoV-19/England/SHEF-CFB1B/2020\|EPI_ISL_613394\|202 | ···················· | ···················· | ···················· | ···················· | ···················· | ···················· |
| O | 6396 | hCoV-19/England/SHEF-CFFE2/2020\|EPI_ISL_613395\|202 | ···················· | ···················· | ···················· | ···················· | ···················· | ···················· |
| O | 6397 | hCoV-19/England/SHEF-D0485/2020\|EPI_ISL_613396\|202 | ···················· | ···················· | ···················· | ···················· | ···················· | ···················· |
| O | 6398 | hCoV-19/England/SHEF-D092C/2020\|EPI_ISL_613397\|202 | ···················· | ···················· | ···················· | ···················· | ···················· | ···················· |
| O | 6399 | hCoV-19/England/SHEF-D094A/2020\|EPI_ISL_613398\|202 | ···················· | ···················· | ···················· | ···················· | ···················· | ···················· |
| O | 6400 | hCoV-19/England/SHEF-D0D02/2020\|EPI_ISL_613399\|202 | ···················· | ···················· | ···················· | ···················· | ···················· | ···················· |
| O | 6401 | hCoV-19/England/ALDP-A87948/2020\|EPI_ISL_606100\|20 | ···················· | ···················· | ···················· | ···················· | ···················· | ···················· |
| O | 6402 | hCoV-19/England/ALDP-A8627B/2020\|EPI_ISL_606101\|20 | ···················· | ···················· | ···················· | ···················· | ···················· | ···················· |
| O | 6403 | hCoV-19/England/ALDP-A86DD4/2020\|EPI_ISL_606102\|20 | ···················· | ···················· | ···················· | ···················· | ···················· | ···················· |
| O | 6404 | hCoV-19/England/ALDP-A87131/2020\|EPI_ISL_606103\|20 | ···················· | ···················· | ···················· | ···················· | ···················· | ···················· |
| O | 6405 | hCoV-19/England/ALDP-A86D10/2020\|EPI_ISL_606104\|20 | ···················· | ···················· | ···················· | ···················· | ···················· | ···················· |
| O | 6406 | hCoV-19/England/ALDP-A87B24/2020\|EPI_ISL_606106\|20 | ···················· | ···················· | ···················· | ···················· | ···················· | ···················· |
| O | 6407 | hCoV-19/England/ALDP-A8783C/2020\|EPI_ISL_606107\|20 | ···················· | ···················· | ···················· | ···················· | ···················· | ···················· |
| O | 6408 | hCoV-19/England/ALDP-A87201/2020\|EPI_ISL_606108\|20 | ···················· | ···················· | ···················· | ···················· | ···················· | ···················· |
| O | 6409 | hCoV-19/England/ALDP-A86F29/2020\|EPI_ISL_606109\|20 | ···················· | ···················· | ···················· | ···················· | ···················· | ···················· |
| O | 6410 | hCoV-19/England/ALDP-A8642A/2020\|EPI_ISL_606111\|20 | ···················· | ···················· | ···················· | ···················· | ···················· | ···················· |

| | | | | | | | | |
|---|---|---|---|---|---|---|---|---|
| O | 6411 | hCoV-19/England/ALDP-A87C8B/2020\|EPI_ISL_606113\|20 | ···················· | ···················· | ···················· | ···················· | ···················· | ···················· |
| O | 6412 | hCoV-19/England/ALDP-A87429/2020\|EPI_ISL_606114\|20 | ···················· | ···················· | ···················· | ···················· | ···················· | ···················· |
| O | 6413 | hCoV-19/England/ALDP-A87070/2020\|EPI_ISL_606115\|20 | ···················· | ···················· | ···················· | ···················· | ···················· | ···················· |
| O | 6414 | hCoV-19/Wales/ALDP-A87632/2020\|EPI_ISL_606116\|2020 | ···················· | ···················· | ···················· | ···················· | ···················· | ···················· |
| O | 6415 | hCoV-19/England/ALDP-A864C0/2020\|EPI_ISL_606118\|20 | ···················· | ···················· | ···················· | ···················· | ···················· | ···················· |
| O | 6416 | hCoV-19/England/ALDP-A87508/2020\|EPI_ISL_606119\|20 | ···················· | ···················· | ···················· | ···················· | ···················· | ···················· |
| O | 6417 | hCoV-19/England/ALDP-A86211/2020\|EPI_ISL_606120\|20 | ···················· | ···················· | ···················· | ···················· | ···················· | ···················· |
| O | 6418 | hCoV-19/England/ALDP-A86D4D/2020\|EPI_ISL_606121\|20 | ···················· | ···················· | ···················· | ···················· | ···················· | ···················· |
| O | 6419 | hCoV-19/England/ALDP-A86EA4/2020\|EPI_ISL_606122\|20 | ···················· | ···················· | ···················· | ···················· | ···················· | ···················· |
| O | 6420 | hCoV-19/England/ALDP-A86220/2020\|EPI_ISL_606123\|20 | ···················· | ···················· | ···················· | ···················· | ···················· | ···················· |
| O | 6421 | hCoV-19/England/ALDP-A86053/2020\|EPI_ISL_606124\|20 | ···················· | ···················· | ···················· | ···················· | ···················· | ···················· |
| O | 6422 | hCoV-19/England/ALDP-A87887/2020\|EPI_ISL_606125\|20 | ···················· | ···················· | ···················· | ···················· | ···················· | ···················· |
| O | 6423 | hCoV-19/England/ALDP-A8775D/2020\|EPI_ISL_606126\|20 | ···················· | ·A·················· | ···················· | ···················· | ···················· | ··A················· |
| O | 6424 | hCoV-19/England/ALDP-A863C3/2020\|EPI_ISL_606128\|20 | ···················· | ·········A·········· | ···················· | ···················· | ···················· | ···················· |
| O | 6425 | hCoV-19/England/ALDP-A86E0E/2020\|EPI_ISL_606129\|20 | ···················· | ···················· | ···················· | ···················· | ···················· | ···················· |
| O | 6426 | hCoV-19/England/ALDP-A8626C/2020\|EPI_ISL_606130\|20 | ···················· | ···················· | ···················· | ···················· | ···················· | ···················· |
| O | 6427 | hCoV-19/England/ALDP-A86D2F/2020\|EPI_ISL_606131\|20 | ···················· | ···················· | ···················· | ···················· | ···················· | ···················· |
| O | 6428 | hCoV-19/England/ALDP-A870AD/2020\|EPI_ISL_606132\|20 | ···················· | ···················· | ···················· | ···················· | ···················· | ···················· |
| O | 6429 | hCoV-19/England/ALDP-A87553/2020\|EPI_ISL_606133\|20 | ···················· | ···················· | ···················· | ···················· | ···················· | ···················· |
| O | 6430 | hCoV-19/England/ALDP-A87474/2020\|EPI_ISL_606134\|20 | ···················· | ···················· | ···················· | ···················· | ······T············· | ···················· |
| O | 6431 | hCoV-19/England/ALDP-A879EE/2020\|EPI_ISL_606135\|20 | ···················· | ···················· | ···················· | ···················· | ···················· | ···················· |
| O | 6432 | hCoV-19/England/ALDP-A8617E/2020\|EPI_ISL_606136\|20 | ···················· | ···················· | ···················· | ···················· | ···················· | ···················· |
| O | 6433 | hCoV-19/England/ALDP-A86CC8/2020\|EPI_ISL_606138\|20 | ···················· | ···················· | ···················· | ···················· | ···················· | ···················· |
| O | 6434 | hCoV-19/England/ALDP-A87C03/2020\|EPI_ISL_606163\|20 | ···················· | ···················· | ···················· | ···················· | ···················· | ···················· |
| O | 6435 | hCoV-19/England/ALDP-A875DB/2020\|EPI_ISL_606164\|20 | ···················· | ···················· | ···················· | ···················· | ···················· | ···················· |
| O | 6436 | hCoV-19/England/ALDP-A875F9/2020\|EPI_ISL_606165\|20 | ···················· | ···················· | ···················· | ···················· | ···················· | ···················· |
| O | 6437 | hCoV-19/England/ALDP-A86E2C/2020\|EPI_ISL_606166\|20 | ···················· | ···················· | ···················· | ···················· | ···················· | ···················· |
| O | 6438 | hCoV-19/England/ALDP-A86509/2020\|EPI_ISL_606167\|20 | ···················· | ···················· | ···················· | ···················· | ···················· | ···················· |
| O | 6439 | hCoV-19/England/ALDP-A87DB5/2020\|EPI_ISL_606168\|20 | ···················· | ···················· | ···················· | ···················· | ···················· | ···················· |
| O | 6440 | hCoV-19/England/ALDP-A8776C/2020\|EPI_ISL_606169\|20 | ···················· | ···················· | ···················· | ···················· | ···················· | ···················· |
| O | 6441 | hCoV-19/England/ALDP-A862A8/2020\|EPI_ISL_606170\|20 | ···················· | ···················· | ···················· | ···················· | ···················· | ···················· |
| O | 6442 | hCoV-19/England/ALDP-A8782D/2020\|EPI_ISL_606171\|20 | ···················· | ···················· | ···················· | ···················· | ···················· | ···················· |
| O | 6443 | hCoV-19/England/ALDP-A87605/2020\|EPI_ISL_606172\|20 | ···················· | ···················· | ···················· | ···················· | ···················· | ···················· |
| O | 6444 | hCoV-19/England/ALDP-A87113/2020\|EPI_ISL_606173\|20 | ···················· | ···················· | ···················· | ···················· | ···················· | ···················· |
| O | 6445 | hCoV-19/England/ALDP-A86DF2/2020\|EPI_ISL_606174\|20 | ···················· | ···················· | ···················· | ···················· | ···················· | ···················· |
| O | 6446 | hCoV-19/England/ALDP-A87B33/2020\|EPI_ISL_606175\|20 | ···················· | ···················· | ···················· | ···················· | ···················· | ···················· |
| O | 6447 | hCoV-19/England/ALDP-A873D1/2020\|EPI_ISL_606176\|20 | ···················· | ···················· | ···················· | ···················· | ···················· | ···················· |
| O | 6448 | hCoV-19/England/ALDP-A865FA/2020\|EPI_ISL_606177\|20 | ···················· | ···················· | ···················· | ···················· | ···················· | ···················· |
| O | 6449 | hCoV-19/England/ALDP-A87A18/2020\|EPI_ISL_606178\|20 | ···················· | ···················· | ···················· | ···················· | ···················· | ···················· |
| O | 6450 | hCoV-19/England/ALDP-A87B51/2020\|EPI_ISL_606180\|20 | ···················· | ···················· | ···················· | ···················· | ···················· | ···················· |
| O | 6451 | hCoV-19/England/ALDP-A8732C/2020\|EPI_ISL_606183\|20 | ···················· | ···················· | ···················· | ···················· | ···················· | ···················· |
| O | 6452 | hCoV-19/England/ALDP-A87BE8/2020\|EPI_ISL_606184\|20 | ···················· | ···················· | ···················· | ···················· | ···················· | ···················· |
| O | 6453 | hCoV-19/England/ALDP-A872D4/2020\|EPI_ISL_606186\|20 | ···················· | ···················· | ···················· | ···················· | ···················· | ···················· |
| O | 6454 | hCoV-19/England/ALDP-A87456/2020\|EPI_ISL_606188\|20 | ···················· | ···················· | ···················· | ···················· | ···················· | ···················· |
| O | 6455 | hCoV-19/England/ALDP-A874A1/2020\|EPI_ISL_606192\|20 | ···················· | ···················· | ···················· | ···················· | ···················· | ···················· |
| O | 6456 | hCoV-19/England/ALDP-A86CE6/2020\|EPI_ISL_606193\|20 | ···················· | ···················· | ···················· | ···················· | ···················· | ···················· |
| O | 6457 | hCoV-19/England/ALDP-A8734A/2020\|EPI_ISL_606194\|20 | ···················· | ···················· | ···················· | ···················· | ···················· | ···················· |
| O | 6458 | hCoV-19/England/ALDP-A86E3B/2020\|EPI_ISL_606195\|20 | ···················· | ···················· | ···················· | ···················· | ···················· | ···················· |
| O | 6459 | hCoV-19/England/ALDP-A87B15/2020\|EPI_ISL_606196\|20 | ···················· | ···················· | ···················· | ···················· | ···················· | ···················· |
| O | 6460 | hCoV-19/England/ALDP-A8769C/2020\|EPI_ISL_606197\|20 | ···················· | ···················· | ···················· | ···················· | ···················· | ···················· |
| O | 6461 | hCoV-19/England/ALDP-A86D89/2020\|EPI_ISL_606198\|20 | ···················· | ···················· | ···················· | ···················· | ···················· | ···················· |
| O | 6462 | hCoV-19/England/ALDP-A87B42/2020\|EPI_ISL_606199\|20 | ···················· | ···················· | ···················· | ···················· | ···················· | ···················· |
| O | 6463 | hCoV-19/England/MILK-A7BD2D/2020\|EPI_ISL_607430\|20 | ···················· | ···················· | ···················· | ···················· | ···················· | ···················· |
| O | 6464 | hCoV-19/England/MILK-A7BB6F/2020\|EPI_ISL_607431\|20 | ···················· | ···················· | ···················· | ···················· | ···················· | ···················· |
| O | 6465 | hCoV-19/England/MILK-A7C894/2020\|EPI_ISL_607432\|20 | ···················· | ···················· | ···················· | ···················· | ···················· | ···················· |
| O | 6466 | hCoV-19/England/MILK-A7C80D/2020\|EPI_ISL_607434\|20 | ···················· | ···················· | ···················· | ···················· | ···················· | ···················· |
| O | 6467 | hCoV-19/England/MILK-A7BF27/2020\|EPI_ISL_607435\|20 | ···················· | ·A·················· | ···················· | ···················· | ···················· | ··A················· |
| O | 6468 | hCoV-19/England/MILK-A7BDA5/2020\|EPI_ISL_607436\|20 | ···················· | ·A·················· | ···················· | ···················· | ···················· | ··A················· |
| O | 6469 | hCoV-19/England/MILK-A7B5AD/2020\|EPI_ISL_607437\|20 | ···················· | ···················· | ···················· | ···················· | ···················· | ···················· |
| O | 6470 | hCoV-19/England/MILK-A7BB9C/2020\|EPI_ISL_607438\|20 | ···················· | ···················· | ···················· | ···················· | ···················· | ···················· |
| O | 6471 | hCoV-19/England/MILK-A7CBB9/2020\|EPI_ISL_607439\|20 | ···················· | ···················· | ···················· | ···················· | ···················· | ···················· |
| O | 6472 | hCoV-19/England/MILK-A7C9EC/2020\|EPI_ISL_607440\|20 | ···················· | ···················· | ···················· | ···················· | ···················· | ···················· |
| O | 6473 | hCoV-19/England/MILK-A7B701/2020\|EPI_ISL_607441\|20 | ···················· | ···················· | ···················· | ···················· | ···················· | ···················· |
| O | 6474 | hCoV-19/England/MILK-A7AFDD/2020\|EPI_ISL_607442\|20 | ···················· | ···················· | ···················· | ···················· | ···················· | ···················· |
| O | 6475 | hCoV-19/England/MILK-A7B69B/2020\|EPI_ISL_607443\|20 | ···················· | ···················· | ···················· | ···················· | ···················· | ···················· |
| O | 6476 | hCoV-19/England/MILK-A7B455/2020\|EPI_ISL_607444\|20 | ···················· | ·A·················· | ···················· | ···················· | ···················· | ··A················· |
| O | 6477 | hCoV-19/England/MILK-A7CA9E/2020\|EPI_ISL_607445\|20 | ···················· | ···················· | ···················· | ···················· | ···················· | ···················· |
| O | 6478 | hCoV-19/England/MILK-A7BBE7/2020\|EPI_ISL_607446\|20 | ···················· | ···················· | ···················· | ···················· | ···················· | ···················· |
| O | 6479 | hCoV-19/England/MILK-A7C4DC/2020\|EPI_ISL_607447\|20 | ···················· | ···················· | ···················· | ···················· | ···················· | ···················· |
| O | 6480 | hCoV-19/England/MILK-A7C199/2020\|EPI_ISL_607449\|20 | ···················· | ···················· | ···················· | ···················· | ···················· | ···················· |
| O | 6481 | hCoV-19/England/MILK-A7BABD/2020\|EPI_ISL_607450\|20 | ···················· | ···················· | ···················· | ···················· | ···················· | ···················· |
| O | 6482 | hCoV-19/England/MILK-A7B25B/2020\|EPI_ISL_607451\|20 | ···················· | ···················· | ···················· | ···················· | ···················· | ···················· |
| O | 6483 | hCoV-19/England/MILK-A7AFA0/2020\|EPI_ISL_607452\|20 | ···················· | ···················· | ···················· | ···················· | ···················· | ···················· |
| O | 6484 | hCoV-19/England/MILK-A7BC5D/2020\|EPI_ISL_607453\|20 | ···················· | ···················· | ···················· | ···················· | ······T············· | ···················· |
| O | 6485 | hCoV-19/England/MILK-A7C858/2020\|EPI_ISL_607454\|20 | ···················· | ···················· | ···················· | ···················· | ···················· | ···················· |
| O | 6486 | hCoV-19/England/MILK-A7BA44/2020\|EPI_ISL_607456\|20 | ···················· | ···················· | ···················· | ···················· | ···················· | ···················· |
| O | 6488 | hCoV-19/England/MILK-A7B0CA/2020\|EPI_ISL_607458\|20 | ···················· | ···················· | ···················· | ···················· | ···················· | ···················· |
| O | 6489 | hCoV-19/England/MILK-A7C16C/2020\|EPI_ISL_607459\|20 | ···················· | ···················· | ···················· | ···················· | ···················· | ···················· |
| O | 6490 | hCoV-19/England/MILK-A7BF45/2020\|EPI_ISL_607462\|20 | ···················· | ···················· | ···················· | ···················· | ···················· | ···················· |
| O | 6491 | hCoV-19/England/MILK-A7B956/2020\|EPI_ISL_607463\|20 | ···················· | ···················· | ···················· | ···················· | ···················· | ···················· |

| | | | | | | | | | |
|---|---|---|---|---|---|---|---|---|---|
| O | 6492 | hCoV-19/England/MILK-A7B7E3/2020\|EPI_ISL_607464\|20 | .................... | .................... | .................... | .................... | .....T.............. | .................... | |
| O | 6493 | hCoV-19/England/MILK-A7B561/2020\|EPI_ISL_607466\|20 | .................... | .................... | .................... | .................... | .................... | .................... | |
| O | 6494 | hCoV-19/England/MILK-A7C4BE/2020\|EPI_ISL_607467\|20 | .................... | .................... | .................... | .................... | .................... | .................... | |
| O | 6495 | hCoV-19/England/MILK-A7B9B0/2020\|EPI_ISL_607468\|20 | .................... | .................... | .................... | .................... | .................... | .................... | |
| O | 6496 | hCoV-19/England/MILK-A7B83B/2020\|EPI_ISL_607469\|20 | .................... | .................... | .................... | .................... | .................... | .................... | |
| O | 6497 | hCoV-19/England/MILK-A7BB50/2020\|EPI_ISL_607491\|20 | .................... | .................... | .................... | .................... | .................... | .................... | |
| O | 6498 | hCoV-19/England/MILK-A7B6AA/2020\|EPI_ISL_607494\|20 | .................... | .................... | .................... | .................... | .................... | .................... | |
| O | 6499 | hCoV-19/England/MILK-A7C7E2/2020\|EPI_ISL_607497\|20 | .................... | .................... | .................... | .................... | .................... | .................... | |
| O | 6500 | hCoV-19/England/MILK-A7B5F8/2020\|EPI_ISL_607498\|20 | .................... | .................... | .................... | .................... | .................... | .................... | |
| O | 6501 | hCoV-19/England/MILK-A7B32B/2020\|EPI_ISL_607499\|20 | .................... | .................... | .................... | .................... | .................... | .................... | |
| O | 6502 | hCoV-19/England/MILK-A6A064/2020\|EPI_ISL_608760\|20 | .................... | .................... | .................... | .................... | .................... | .................... | |
| O | 6503 | hCoV-19/England/MILK-A6BA65/2020\|EPI_ISL_608761\|20 | .................... | .................... | .................... | .................... | .................... | .................... | |
| O | 6504 | hCoV-19/England/MILK-A6B485/2020\|EPI_ISL_608762\|20 | .................... | .................... | .................... | .................... | .................... | .................... | |
| O | 6505 | hCoV-19/England/MILK-A6B124/2020\|EPI_ISL_608763\|20 | .................... | .................... | .................... | .................... | .................... | .................... | |
| O | 6506 | hCoV-19/England/MILK-A6A037/2020\|EPI_ISL_608764\|20 | .................... | .................... | .................... | .................... | .................... | .................... | |
| O | 6507 | hCoV-19/England/MILK-A6AEC4/2020\|EPI_ISL_608765\|20 | .................... | .................... | .................... | .................... | .................... | .................... | |
| O | 6508 | hCoV-19/England/MILK-A6B546/2020\|EPI_ISL_608766\|20 | .................... | .................... | .................... | .................... | .................... | .................... | |
| O | 6509 | hCoV-19/England/MILK-A69D22/2020\|EPI_ISL_608767\|20 | .................... | .................... | .................... | .................... | .................... | .................... | |
| O | 6510 | hCoV-19/England/MILK-A6BAFC/2020\|EPI_ISL_608771\|20 | .................... | .................... | .................... | .................... | .................... | .................... | |
| O | 6511 | hCoV-19/England/MILK-A69D31/2020\|EPI_ISL_608772\|20 | .................... | .................... | .................... | .................... | .................... | .................... | |
| O | 6512 | hCoV-19/England/MILK-A69F1D/2020\|EPI_ISL_608773\|20 | .................... | .................... | .................... | .................... | .................... | .................... | |
| O | 6513 | hCoV-19/England/MILK-A6AC06/2020\|EPI_ISL_608775\|20 | .................... | .................... | .................... | .................... | .................... | .................... | |
| O | 6514 | hCoV-19/England/MILK-A69E2F/2020\|EPI_ISL_608776\|20 | .................... | .A.................. | .................... | .................... | .................... | ..A................. | |
| O | 6515 | hCoV-19/England/MILK-A6BA92/2020\|EPI_ISL_608777\|20 | .................... | .................... | .................... | .................... | .................... | .................... | |
| O | 6516 | hCoV-19/England/MILK-A6AC42/2020\|EPI_ISL_608778\|20 | .................... | .................... | .................... | .................... | .................... | .................... | |
| O | 6517 | hCoV-19/England/MILK-A6B25E/2020\|EPI_ISL_608779\|20 | .................... | .................... | .................... | .................... | .................... | .................... | |
| O | 6518 | hCoV-19/England/MILK-A6AF49/2020\|EPI_ISL_608780\|20 | .................... | .................... | .................... | .................... | .................... | .................... | |
| O | 6519 | hCoV-19/England/MILK-A6B573/2020\|EPI_ISL_608782\|20 | .................... | .................... | .................... | .................... | .................... | .................... | |
| O | 6520 | hCoV-19/Wales/MILK-A6B42B/2020\|EPI_ISL_608783\|2020 | .................... | .................... | .................... | .................... | .................... | .................... | |
| O | 6521 | hCoV-19/England/MILK-A6ACCA/2020\|EPI_ISL_608784\|20 | .................... | .................... | .................... | .................... | .................... | .................... | |
| O | 6522 | hCoV-19/England/MILK-A6AE4C/2020\|EPI_ISL_608786\|20 | .................... | .................... | .................... | .................... | .................... | .................... | |
| O | 6523 | hCoV-19/England/MILK-A6B0EB/2020\|EPI_ISL_608788\|20 | .................... | .................... | .................... | .................... | .................... | .................... | |
| O | 6524 | hCoV-19/England/MILK-A6BB80/2020\|EPI_ISL_608789\|20 | .................... | .................... | .................... | .................... | .................... | .................... | |
| O | 6525 | hCoV-19/England/MILK-A6AFA3/2020\|EPI_ISL_608790\|20 | .................... | .................... | .................... | .................... | .................... | .................... | |
| O | 6526 | hCoV-19/England/MILK-A6ACAC/2020\|EPI_ISL_608791\|20 | .................... | .................... | .................... | .................... | .................... | .................... | |
| O | 6527 | hCoV-19/England/MILK-A6B43A/2020\|EPI_ISL_608792\|20 | .................... | .................... | .................... | .................... | .................... | .................... | |
| O | 6528 | hCoV-19/England/MILK-A69FA4/2020\|EPI_ISL_608793\|20 | .................... | .................... | .................... | .................... | .................... | .................... | |
| O | 6529 | hCoV-19/England/MILK-A6AFFE/2020\|EPI_ISL_608794\|20 | .................... | .................... | .................... | .................... | .................... | .................... | |
| O | 6530 | hCoV-19/England/MILK-A6B4FE/2020\|EPI_ISL_608795\|20 | .................... | .................... | .................... | .................... | .................... | .................... | |
| O | 6531 | hCoV-19/England/MILK-A6A161/2020\|EPI_ISL_608797\|20 | .................... | .................... | .................... | .................... | .................... | .................... | |
| O | 6532 | hCoV-19/England/PORT-2F5770/2020\|EPI_ISL_613300\|20 | .................... | .................... | .................... | .................... | .................... | .................... | |
| O | 6533 | hCoV-19/England/PORT-2F57DA/2020\|EPI_ISL_613301\|20 | .................... | .................... | .................... | .................... | .................... | .................... | |
| O | 6534 | hCoV-19/England/PORT-2F5E3F/2020\|EPI_ISL_613302\|20 | .................... | .................... | .................... | .................... | .................... | .................... | |
| O | 6535 | hCoV-19/England/PORT-2F5E4E/2020\|EPI_ISL_613303\|20 | .................... | .................... | .................... | .................... | .................... | .................... | |
| O | 6536 | hCoV-19/England/PORT-2F5E5D/2020\|EPI_ISL_613304\|20 | .................... | .................... | .................... | .................... | .................... | .................... | |
| O | 6537 | hCoV-19/England/PORT-2F5E7B/2020\|EPI_ISL_613305\|20 | .................... | .................... | .................... | .................... | .................... | .................... | |
| O | 6538 | hCoV-19/England/PORT-2F5EA8/2020\|EPI_ISL_613306\|20 | .................... | .................... | .................... | .................... | .................... | .................... | |
| P | 1 | hCoV-19/Gibraltar/204462836/2020\|EPI_ISL_615010\|20 | .................... | .................... | .................... | .................... | .................... | .................... | |
| P | 2 | hCoV-19/Gibraltar/204462838/2020\|EPI_ISL_615011\|20 | .................... | .................... | .................... | .................... | .................... | .................... | |
| P | 3 | hCoV-19/Gibraltar/204462840/2020\|EPI_ISL_615012\|20 | .................... | .................... | .................... | .................... | .................... | .................... | |
| P | 4 | hCoV-19/Gibraltar/204462841/2020\|EPI_ISL_615013\|20 | .................... | .................... | .................... | .................... | .....T.............. | .................... | |
| P | 5 | hCoV-19/Gibraltar/204462843/2020\|EPI_ISL_615014\|20 | .................... | .................... | .................... | .................... | .................... | .................... | |
| P | 6 | hCoV-19/Gibraltar/204462844/2020\|EPI_ISL_615015\|20 | .................... | .................... | .................... | .................... | .................... | .................... | |
| P | 7 | hCoV-19/Gibraltar/204462846/2020\|EPI_ISL_615016\|20 | .................... | .................... | .................... | .................... | .................... | .................... | |
| P | 8 | hCoV-19/Gibraltar/204462847/2020\|EPI_ISL_615017\|20 | .................... | .................... | .................... | .................... | .................... | .................... | |
| P | 9 | hCoV-19/Gibraltar/204462848/2020\|EPI_ISL_615018\|20 | .................... | .................... | .................... | .................... | .................... | .................... | |
| P | 10 | hCoV-19/Gibraltar/204462849/2020\|EPI_ISL_615019\|20 | .................... | .................... | .................... | .................... | .................... | .................... | |
| P | 11 | hCoV-19/Gibraltar/204462850/2020\|EPI_ISL_615020\|20 | .................... | .................... | .................... | .................... | .................... | .................... | |
| P | 12 | hCoV-19/Gibraltar/204462851/2020\|EPI_ISL_615021\|20 | .................... | .................... | .................... | .................... | .................... | .................... | |
| P | 13 | hCoV-19/Gibraltar/204462855/2020\|EPI_ISL_615023\|20 | .................... | .................... | .................... | .................... | .................... | .................... | |
| P | 14 | hCoV-19/Gibraltar/204462856/2020\|EPI_ISL_615024\|20 | .................... | .................... | .................... | .................... | .................... | .................... | |
| P | 15 | hCoV-19/Gibraltar/204462857/2020\|EPI_ISL_615025\|20 | .................... | .................... | .................... | .................... | .................... | .................... | |
| P | 16 | hCoV-19/Gibraltar/204462858/2020\|EPI_ISL_615026\|20 | ..................M. | .................... | .................... | .................... | .................... | .................... | 1 |
| P | 17 | hCoV-19/Gibraltar/204462859/2020\|EPI_ISL_615027\|20 | .................... | .................... | .................... | .................... | .................... | .................... | |
| P | 18 | hCoV-19/Gibraltar/204462860/2020\|EPI_ISL_615028\|20 | .................... | .................... | .................... | .................... | .................... | .................... | |
| P | 19 | hCoV-19/Gibraltar/204462862/2020\|EPI_ISL_615029\|20 | .................... | .................... | .................... | .................... | .................... | .................... | |
| P | 20 | hCoV-19/Gibraltar/204462863/2020\|EPI_ISL_615030\|20 | .................... | .................... | .................... | .................... | .................... | .................... | |
| P | 21 | hCoV-19/Gibraltar/204462864/2020\|EPI_ISL_615031\|20 | .................... | .................... | .................... | .................... | .................... | .................... | |
| P | 22 | hCoV-19/Gibraltar/204462865/2020\|EPI_ISL_615032\|20 | .................... | .................... | .................... | .................... | .................... | .................... | |
| P | 23 | hCoV-19/Gibraltar/204462866/2020\|EPI_ISL_615033\|20 | .................... | .................... | .................... | .................... | .....T.............. | .................... | |
| P | 24 | hCoV-19/Gibraltar/204462867/2020\|EPI_ISL_615034\|20 | .................... | .................... | .................... | .................... | .................... | .................... | |
| P | 25 | hCoV-19/Gibraltar/204462868/2020\|EPI_ISL_615035\|20 | .................... | .................... | .................... | .................... | .................... | .................... | |
| P | 26 | hCoV-19/Gibraltar/204462870/2020\|EPI_ISL_615037\|20 | .................... | .................... | .................... | .................... | .................... | .................... | |
| P | 27 | hCoV-19/Gibraltar/204462871/2020\|EPI_ISL_615038\|20 | .................... | .................... | .................... | .................... | .................... | .................... | |
| P | 28 | hCoV-19/Gibraltar/204462872/2020\|EPI_ISL_615039\|20 | .................... | .................... | .................... | .................... | .................... | .................... | |
| P | 29 | hCoV-19/Gibraltar/204462873/2020\|EPI_ISL_615040\|20 | .................... | .................... | .................... | .................... | .................... | .................... | |
| P | 30 | hCoV-19/Gibraltar/204462874/2020\|EPI_ISL_615041\|20 | .................... | .................... | .................... | .................... | .....T.............. | .................... | |
| P | 31 | hCoV-19/Gibraltar/204462875/2020\|EPI_ISL_615042\|20 | .................... | .................... | .................... | .................... | .................... | .................... | |
| P | 32 | hCoV-19/Gibraltar/204462877/2020\|EPI_ISL_615044\|20 | .................... | .................... | .................... | .................... | .................... | .................... | |
| P | 33 | hCoV-19/Gibraltar/204462878/2020\|EPI_ISL_615045\|20 | .................... | .................... | .................... | .................... | .................... | .................... | |

| | | | | | | | | |
|---|---|---|---|---|---|---|---|---|
| P | 34 | hCoV-19/Gibraltar/204462879/2020\|EPI_ISL_615046\|20 | ···················· | ···················· | ····················· | ···················· | ···················· | ···················· |
| P | 35 | hCoV-19/Gibraltar/204462880/2020\|EPI_ISL_615047\|20 | ···················· | ···················· | ····················· | ···················· | ···················· | ···················· |
| P | 36 | hCoV-19/Gibraltar/204462881/2020\|EPI_ISL_615048\|20 | ···················· | ···················· | ····················· | ···················· | ···················· | ···················· |
| P | 37 | hCoV-19/Gibraltar/204462882/2020\|EPI_ISL_615049\|20 | ···················· | ···················· | ····················· | ···················· | ···················· | ···················· |
| P | 38 | hCoV-19/Denmark/DCGC-3511/2020\|EPI_ISL_616300\|2020 | ···················· | ···················· | ····················· | ···················· | ···················· | ···················· |
| P | 39 | hCoV-19/Denmark/DCGC-3512/2020\|EPI_ISL_616301\|2020 | ···················· | ···················· | ····················· | ···················· | ···················· | ···················· |
| P | 40 | hCoV-19/Denmark/DCGC-3271/2020\|EPI_ISL_616303\|2020 | ···················· | ···················· | ····················· | ···················· | ···················· | ···················· |
| P | 41 | hCoV-19/Denmark/DCGC-3272/2020\|EPI_ISL_616304\|2020 | ···················· | ···················· | ····················· | ···················· | ···················· | ···················· |
| P | 42 | hCoV-19/Denmark/DCGC-3514/2020\|EPI_ISL_616305\|2020 | ···················· | ···················· | ····················· | ···················· | ···················· | ···················· |
| P | 43 | hCoV-19/Denmark/DCGC-3515/2020\|EPI_ISL_616306\|2020 | ···················· | ···················· | ····················· | ···················· | ···················· | ···················· |
| P | 44 | hCoV-19/Denmark/DCGC-3516/2020\|EPI_ISL_616307\|2020 | ···················· | ···················· | ····················· | ···················· | ···················· | ···················· |
| P | 45 | hCoV-19/Denmark/DCGC-3517/2020\|EPI_ISL_616308\|2020 | ·······T············ | ···················· | ····················· | ···················· | ···················· | ···················· |
| P | 46 | hCoV-19/Denmark/DCGC-3518/2020\|EPI_ISL_616309\|2020 | ···················· | ···················· | ····················· | ···················· | ···················· | ···················· |
| P | 47 | hCoV-19/Denmark/DCGC-3523/2020\|EPI_ISL_616314\|2020 | ···················· | ···················· | ····················· | ···················· | ···················· | ···················· |
| P | 48 | hCoV-19/Denmark/DCGC-3524/2020\|EPI_ISL_616315\|2020 | ···················· | ···················· | ····················· | ···················· | ···················· | ···················· |
| P | 49 | hCoV-19/Denmark/DCGC-3525/2020\|EPI_ISL_616316\|2020 | ···················· | ···················· | ····················· | ···················· | ···················· | ···················· |
| P | 50 | hCoV-19/Denmark/DCGC-3526/2020\|EPI_ISL_616317\|2020 | ···················· | ···················· | ····················· | ···················· | ···················· | ···················· |
| P | 51 | hCoV-19/Denmark/DCGC-3527/2020\|EPI_ISL_616318\|2020 | ···················· | ···················· | ····················· | ···················· | ···················· | ···················· |
| P | 52 | hCoV-19/Denmark/DCGC-3528/2020\|EPI_ISL_616319\|2020 | ···················· | ···················· | ····················· | ···················· | ···················· | ···················· |
| P | 53 | hCoV-19/Denmark/DCGC-3547/2020\|EPI_ISL_616340\|2020 | ···················· | ·A·················· | ····················· | ···················· | ···················· | ··A················· |
| P | 54 | hCoV-19/Denmark/DCGC-3548/2020\|EPI_ISL_616341\|2020 | ···················· | ···················· | ····················· | ···················· | ···················· | ···················· |
| P | 55 | hCoV-19/Denmark/DCGC-3549/2020\|EPI_ISL_616342\|2020 | ···················· | ···················· | ····················· | ···················· | ···················· | ···················· |
| P | 56 | hCoV-19/Denmark/DCGC-3550/2020\|EPI_ISL_616343\|2020 | ···················· | ···················· | ····················· | ···················· | ···················· | ···················· |
| P | 57 | hCoV-19/Denmark/DCGC-3552/2020\|EPI_ISL_616345\|2020 | ···················· | ···················· | ····················· | ···················· | ···················· | ···················· |
| P | 58 | hCoV-19/Denmark/DCGC-3553/2020\|EPI_ISL_616346\|2020 | ···················· | ···················· | ····················· | ···················· | ···················· | ···················· |
| P | 59 | hCoV-19/Denmark/DCGC-3554/2020\|EPI_ISL_616347\|2020 | ···················· | ···················· | ····················· | ···················· | ···················· | ···················· |
| P | 60 | hCoV-19/Denmark/DCGC-3555/2020\|EPI_ISL_616348\|2020 | ···················· | ···················· | ····················· | ···················· | ···················· | ···················· |
| P | 61 | hCoV-19/Denmark/DCGC-3556/2020\|EPI_ISL_616349\|2020 | ···················· | ·A·················· | ····················· | ···················· | ···················· | ··A················· |
| P | 62 | hCoV-19/Denmark/DCGC-3275/2020\|EPI_ISL_616350\|2020 | ···················· | ···················· | ····················· | ···················· | ···················· | ···················· |
| P | 63 | hCoV-19/Denmark/DCGC-3557/2020\|EPI_ISL_616351\|2020 | ···················· | ···················· | ····················· | ···················· | ···················· | ···················· |
| P | 64 | hCoV-19/Denmark/DCGC-3276/2020\|EPI_ISL_616352\|2020 | ···················· | ···················· | ····················· | ···················· | ···················· | ···················· |
| P | 65 | hCoV-19/Denmark/DCGC-3558/2020\|EPI_ISL_616353\|2020 | ···················· | ···················· | ····················· | ···················· | ···················· | ···················· |
| P | 66 | hCoV-19/Denmark/DCGC-3559/2020\|EPI_ISL_616354\|2020 | ···················· | ···················· | ····················· | ···················· | ···················· | ···················· |
| P | 67 | hCoV-19/Denmark/DCGC-3560/2020\|EPI_ISL_616355\|2020 | ···················· | ···················· | ····················· | ···················· | ···················· | ···················· |
| P | 68 | hCoV-19/Denmark/DCGC-3561/2020\|EPI_ISL_616356\|2020 | ···················· | ···················· | ····················· | ···················· | ···················· | ···················· |
| P | 69 | hCoV-19/Denmark/DCGC-3563/2020\|EPI_ISL_616358\|2020 | ···················· | ···················· | ····················· | ···················· | ···················· | ···················· |
| P | 70 | hCoV-19/Denmark/DCGC-3564/2020\|EPI_ISL_616359\|2020 | ···················· | ···················· | ····················· | ···················· | ···················· | ···················· |
| P | 71 | hCoV-19/Denmark/DCGC-3565/2020\|EPI_ISL_616360\|2020 | ···················· | ···················· | ····················· | ···················· | ···················· | ···················· |
| P | 72 | hCoV-19/Denmark/DCGC-3566/2020\|EPI_ISL_616361\|2020 | ···················· | ···················· | ····················· | ···················· | ···················· | ···················· |
| P | 73 | hCoV-19/Denmark/DCGC-3567/2020\|EPI_ISL_616362\|2020 | ···················· | ···················· | ····················· | ···················· | ···················· | ···················· |
| P | 74 | hCoV-19/Denmark/DCGC-3568/2020\|EPI_ISL_616363\|2020 | ···················· | ···················· | ····················· | ···················· | ···················· | ···················· |
| P | 75 | hCoV-19/Denmark/DCGC-3569/2020\|EPI_ISL_616364\|2020 | ···················· | ···················· | ····················· | ···················· | ···················· | ···················· |
| P | 76 | hCoV-19/Denmark/DCGC-3570/2020\|EPI_ISL_616365\|2020 | ···················· | ···················· | ····················· | ···················· | ···················· | ···················· |
| P | 77 | hCoV-19/Denmark/DCGC-3571/2020\|EPI_ISL_616366\|2020 | ···················· | ···················· | ····················· | ···················· | ···················· | ···················· |
| P | 78 | hCoV-19/Denmark/DCGC-3572/2020\|EPI_ISL_616367\|2020 | ···················· | ···················· | ····················· | ···················· | ···················· | ···················· |
| P | 79 | hCoV-19/Denmark/DCGC-3277/2020\|EPI_ISL_616368\|2020 | ···················· | ···················· | ····················· | ···················· | ···················· | ···················· |
| P | 80 | hCoV-19/Denmark/DCGC-3279/2020\|EPI_ISL_616370\|2020 | ···················· | ···················· | ····················· | ···················· | ···················· | ···················· |
| P | 81 | hCoV-19/Denmark/DCGC-3605/2020\|EPI_ISL_616371\|2020 | ···················· | ···················· | ····················· | ···················· | ···················· | ···················· |
| P | 82 | hCoV-19/Denmark/DCGC-3606/2020\|EPI_ISL_616372\|2020 | ···················· | ···················· | ····················· | ···················· | ···················· | ···················· |
| P | 83 | hCoV-19/Denmark/DCGC-3607/2020\|EPI_ISL_616373\|2020 | ···················· | ·A·················· | ····················· | ···················· | ···················· | ··A················· |
| P | 84 | hCoV-19/Denmark/DCGC-3608/2020\|EPI_ISL_616374\|2020 | ···················· | ···················· | ····················· | ···················· | ···················· | ···················· |
| P | 85 | hCoV-19/Denmark/DCGC-3609/2020\|EPI_ISL_616375\|2020 | ···················· | ···················· | ····················· | ···················· | ···················· | ···················· |
| P | 86 | hCoV-19/Denmark/DCGC-3610/2020\|EPI_ISL_616376\|2020 | ···················· | ···················· | ····················· | ···················· | ···················· | ···················· |
| P | 87 | hCoV-19/Denmark/DCGC-6298/2020\|EPI_ISL_616377\|2020 | ···················· | ···················· | ····················· | ···················· | ···················· | ···················· |
| P | 88 | hCoV-19/Denmark/DCGC-3611/2020\|EPI_ISL_616378\|2020 | ···················· | ···················· | ····················· | ···················· | ···················· | ···················· |
| P | 89 | hCoV-19/Denmark/DCGC-3612/2020\|EPI_ISL_616379\|2020 | ···················· | ···················· | ····················· | ···················· | ···················· | ···················· |
| P | 90 | hCoV-19/Denmark/DCGC-2418/2020\|EPI_ISL_617600\|2020 | ···················· | ···················· | ····················· | ···················· | ···················· | ···················· |
| P | 91 | hCoV-19/Denmark/DCGC-2419/2020\|EPI_ISL_617601\|2020 | ···················· | ···················· | ····················· | ···················· | ···················· | ···················· |
| P | 92 | hCoV-19/Denmark/DCGC-2420/2020\|EPI_ISL_617602\|2020 | ···················· | ···················· | ····················· | ···················· | ···················· | ···················· |
| P | 93 | hCoV-19/Denmark/DCGC-2421/2020\|EPI_ISL_617603\|2020 | ···················· | ···················· | ····················· | ···················· | ···················· | ···················· |
| P | 94 | hCoV-19/Denmark/DCGC-2429/2020\|EPI_ISL_617604\|2020 | ···················· | ···················· | ····················· | ···················· | ···················· | ···················· |
| P | 95 | hCoV-19/Denmark/DCGC-2430/2020\|EPI_ISL_617605\|2020 | ···················· | ···················· | ····················· | ···················· | ···················· | ···················· |
| P | 96 | hCoV-19/Denmark/DCGC-2431/2020\|EPI_ISL_617606\|2020 | ···················· | ···················· | ····················· | ···················· | ···················· | ···················· |
| P | 97 | hCoV-19/Denmark/DCGC-2432/2020\|EPI_ISL_617607\|2020 | ···················· | ···················· | ····················· | ···················· | ···················· | ···················· |
| P | 98 | hCoV-19/Denmark/DCGC-2433/2020\|EPI_ISL_617608\|2020 | ···················· | ···················· | ····················· | ···················· | ···················· | ···················· |
| P | 99 | hCoV-19/Denmark/DCGC-2434/2020\|EPI_ISL_617609\|2020 | ···················· | ···················· | ····················· | ···················· | ···················· | ···················· |
| P | 100 | hCoV-19/Denmark/DCGC-2435/2020\|EPI_ISL_617610\|2020 | ···················· | ···················· | ····················· | ···················· | ···················· | ···················· |
| P | 101 | hCoV-19/Denmark/DCGC-2436/2020\|EPI_ISL_617611\|2020 | ···················· | ···················· | ····················· | ···················· | ···················· | ···················· |
| P | 102 | hCoV-19/Denmark/DCGC-2437/2020\|EPI_ISL_617612\|2020 | ···················· | ···················· | ····················· | ···················· | ···················· | ···················· |
| P | 103 | hCoV-19/Denmark/DCGC-2438/2020\|EPI_ISL_617613\|2020 | ···················· | ···················· | ····················· | ···················· | ···················· | ···················· |
| P | 104 | hCoV-19/Denmark/DCGC-2439/2020\|EPI_ISL_617614\|2020 | ···················· | ···················· | ····················· | ···················· | ···················· | ···················· |
| P | 105 | hCoV-19/Denmark/DCGC-2440/2020\|EPI_ISL_617615\|2020 | ···················· | ···················· | ····················· | ···················· | ···················· | ···················· |
| P | 106 | hCoV-19/Denmark/DCGC-2441/2020\|EPI_ISL_617616\|2020 | ···················· | ···················· | ····················· | ···················· | ···················· | ···················· |
| P | 107 | hCoV-19/Denmark/DCGC-2442/2020\|EPI_ISL_617617\|2020 | ···················· | ·A·················· | ····················· | ···················· | ···················· | ···················· |
| P | 108 | hCoV-19/Denmark/DCGC-2262/2020\|EPI_ISL_617619\|2020 | ···················· | ···················· | ····················· | ···················· | ···················· | ···················· |
| P | 109 | hCoV-19/Denmark/DCGC-2263/2020\|EPI_ISL_617620\|2020 | ···················· | ···················· | ····················· | ···················· | ···················· | ···················· |
| P | 110 | hCoV-19/Denmark/DCGC-2264/2020\|EPI_ISL_617621\|2020 | ···················· | ···················· | ····················· | ···················· | ···················· | ···················· |
| P | 111 | hCoV-19/Denmark/DCGC-2265/2020\|EPI_ISL_617622\|2020 | ···················· | ···················· | ····················· | ···················· | ···················· | ···················· |
| P | 112 | hCoV-19/Denmark/DCGC-2266/2020\|EPI_ISL_617623\|2020 | ···················· | ···················· | ····················· | ···················· | ···················· | ···················· |
| P | 113 | hCoV-19/Denmark/DCGC-2267/2020\|EPI_ISL_617624\|2020 | ···················· | ··················C· | ····················· | ···················· | ···················· | ···················· 1 |

| | | | | | | | |
|---|---|---|---|---|---|---|---|
| P | 114 hCoV-19/Denmark/DCGC-2268/2020\|EPI_ISL_617625\|2020 | ···················· | ···················· | ····················· | ···················· | ···················· | ···················· |
| P | 115 hCoV-19/Denmark/DCGC-2269/2020\|EPI_ISL_617626\|2020 | ···················· | ···················· | ····················· | ···················· | ···················· | ···················· |
| P | 116 hCoV-19/Denmark/DCGC-2270/2020\|EPI_ISL_617627\|2020 | ···················· | ···················· | ····················· | ···················· | ···················· | ···················· |
| P | 117 hCoV-19/Denmark/DCGC-2271/2020\|EPI_ISL_617628\|2020 | ···················· | ···················· | ····················· | ···················· | ···················· | ···················· |
| P | 118 hCoV-19/Denmark/DCGC-2272/2020\|EPI_ISL_617629\|2020 | ···················· | ···················· | ····················· | ···················· | ···················· | ···················· |
| P | 119 hCoV-19/Denmark/DCGC-2273/2020\|EPI_ISL_617630\|2020 | ···················· | ···················· | ····················· | ···················· | ···················· | ···················· |
| P | 120 hCoV-19/Denmark/DCGC-2274/2020\|EPI_ISL_617631\|2020 | ···················· | ···················· | ····················· | ···················· | ···················· | ···················· |
| P | 121 hCoV-19/Denmark/DCGC-2275/2020\|EPI_ISL_617632\|2020 | ···················· | ···················· | ····················· | ···················· | ···················· | ···················· |
| P | 122 hCoV-19/Denmark/DCGC-2684/2020\|EPI_ISL_617633\|2020 | ···················· | ·A·················· | ····················· | ···················· | ···················· | ···················· |
| P | 123 hCoV-19/Denmark/DCGC-2276/2020\|EPI_ISL_617634\|2020 | ···················· | ·A·················· | ····················· | ···················· | ···················· | ··A················· |
| P | 124 hCoV-19/Denmark/DCGC-2277/2020\|EPI_ISL_617635\|2020 | ···················· | ···················· | ····················· | ···················· | ···················· | ···················· |
| P | 125 hCoV-19/Denmark/DCGC-2278/2020\|EPI_ISL_617636\|2020 | ···················· | ···················· | ····················· | ···················· | ···················· | ···················· |
| P | 126 hCoV-19/Denmark/DCGC-2279/2020\|EPI_ISL_617637\|2020 | ···················· | ···················· | ····················· | ···················· | ···················· | ···················· |
| P | 127 hCoV-19/Denmark/DCGC-2281/2020\|EPI_ISL_617639\|2020 | ···················· | ···················· | ····················· | ···················· | ···················· | ···················· |
| P | 128 hCoV-19/Denmark/DCGC-2282/2020\|EPI_ISL_617640\|2020 | ···················· | ···················· | ····················· | ···················· | ···················· | ···················· |
| P | 129 hCoV-19/Denmark/DCGC-2286/2020\|EPI_ISL_617644\|2020 | ···················· | ···················· | ····················· | ···················· | ···················· | ···················· |
| P | 130 hCoV-19/Denmark/DCGC-2287/2020\|EPI_ISL_617645\|2020 | ···················· | ···················· | ····················· | ···················· | ···················· | ···················· |
| P | 131 hCoV-19/Denmark/DCGC-2288/2020\|EPI_ISL_617646\|2020 | ···················· | ···················· | ····················· | ···················· | ···················· | ···················· |
| P | 132 hCoV-19/Denmark/DCGC-2289/2020\|EPI_ISL_617647\|2020 | ···················· | ···················· | ····················· | ···················· | ···················· | ···················· |
| P | 133 hCoV-19/Denmark/DCGC-2290/2020\|EPI_ISL_617648\|2020 | ···················· | ···················· | ····················· | ···················· | ···················· | ···················· |
| P | 134 hCoV-19/Denmark/DCGC-2291/2020\|EPI_ISL_617649\|2020 | ···················· | ···················· | ····················· | ···················· | ···················· | ···················· |
| P | 135 hCoV-19/Denmark/DCGC-2332/2020\|EPI_ISL_617670\|2020 | ···················· | ···················· | ····················· | ···················· | ···················· | ···················· |
| P | 136 hCoV-19/Denmark/DCGC-2333/2020\|EPI_ISL_617671\|2020 | ···················· | ···················· | ····················· | ···················· | ···················· | ···················· |
| P | 137 hCoV-19/Denmark/DCGC-2334/2020\|EPI_ISL_617672\|2020 | ···················· | ···················· | ····················· | ···················· | ···················· | ···················· |
| P | 138 hCoV-19/Denmark/DCGC-2335/2020\|EPI_ISL_617673\|2020 | ···················· | ···················· | ····················· | ···················· | ···················· | ···················· |
| P | 139 hCoV-19/Denmark/DCGC-2336/2020\|EPI_ISL_617674\|2020 | ···················· | ···················· | ····················· | ···················· | ···················· | ···················· |
| P | 140 hCoV-19/Denmark/DCGC-2337/2020\|EPI_ISL_617675\|2020 | ···················· | ···················· | ····················· | ···················· | ···················· | ···················· |
| P | 141 hCoV-19/Denmark/DCGC-2338/2020\|EPI_ISL_617676\|2020 | ···················· | ···················· | ····················· | ···················· | ···················· | ···················· |
| P | 142 hCoV-19/Denmark/DCGC-2339/2020\|EPI_ISL_617677\|2020 | ···················· | ···················· | ····················· | ···················· | ···················· | ···················· |
| P | 143 hCoV-19/Denmark/DCGC-2340/2020\|EPI_ISL_617678\|2020 | ···················· | ···················· | ····················· | ···················· | ···················· | ···················· |
| P | 144 hCoV-19/Denmark/DCGC-2341/2020\|EPI_ISL_617679\|2020 | ···················· | ···················· | ····················· | ···················· | ···················· | ···················· |
| P | 145 hCoV-19/Denmark/DCGC-2342/2020\|EPI_ISL_617680\|2020 | ···················· | ···················· | ····················· | ···················· | ···················· | ···················· |
| P | 146 hCoV-19/Denmark/DCGC-2343/2020\|EPI_ISL_617681\|2020 | ···················· | ···················· | ····················· | ···················· | ···················· | ···················· |
| P | 147 hCoV-19/Denmark/DCGC-2344/2020\|EPI_ISL_617682\|2020 | ···················· | ···················· | ····················· | ···················· | ···················· | ···················· |
| P | 148 hCoV-19/Denmark/DCGC-2345/2020\|EPI_ISL_617683\|2020 | ···················· | ···················· | ····················· | ···················· | ···················· | ···················· |
| P | 149 hCoV-19/Denmark/DCGC-2346/2020\|EPI_ISL_617684\|2020 | ···················· | ···················· | ····················· | ···················· | ···················· | ···················· |
| P | 150 hCoV-19/Denmark/DCGC-2347/2020\|EPI_ISL_617685\|2020 | ···················· | ···················· | ····················· | ···················· | ···················· | ···················· |
| P | 151 hCoV-19/Denmark/DCGC-2348/2020\|EPI_ISL_617686\|2020 | ···················· | ···················· | ····················· | ···················· | ···················· | ···················· |
| P | 152 hCoV-19/Denmark/DCGC-2349/2020\|EPI_ISL_617687\|2020 | ···················· | ···················· | ····················· | ···················· | ···················· | ···················· |
| P | 153 hCoV-19/Denmark/DCGC-2350/2020\|EPI_ISL_617688\|2020 | ···················· | ···················· | ····················· | ···················· | ···················· | ···················· |
| P | 154 hCoV-19/Denmark/DCGC-2351/2020\|EPI_ISL_617689\|2020 | ···················· | ···················· | ····················· | ···················· | ···················· | ···················· |
| P | 155 hCoV-19/Denmark/DCGC-2352/2020\|EPI_ISL_617690\|2020 | ···················· | ···················· | ····················· | ···················· | ···················· | ···················· |
| P | 156 hCoV-19/Denmark/DCGC-2353/2020\|EPI_ISL_617691\|2020 | ···················· | ···················· | ····················· | ···················· | ···················· | ···················· |
| P | 157 hCoV-19/Denmark/DCGC-2354/2020\|EPI_ISL_617692\|2020 | ···················· | ···················· | ····················· | ···················· | ···················· | ···················· |
| P | 158 hCoV-19/Denmark/DCGC-2355/2020\|EPI_ISL_617693\|2020 | ···················· | ···················· | ····················· | ···················· | ···················· | ···················· |
| P | 159 hCoV-19/Denmark/DCGC-2690/2020\|EPI_ISL_617694\|2020 | ···················· | ···················· | ····················· | ···················· | ···················· | ···················· |
| P | 160 hCoV-19/Denmark/DCGC-2356/2020\|EPI_ISL_617695\|2020 | ···················· | ···················· | ····················· | ···················· | ···················· | ···················· |
| P | 161 hCoV-19/Denmark/DCGC-2357/2020\|EPI_ISL_617696\|2020 | ···················· | ···················· | ····················· | ···················· | ···················· | ···················· |
| P | 162 hCoV-19/Denmark/DCGC-2358/2020\|EPI_ISL_617698\|2020 | ···················· | ···················· | ····················· | ···················· | ···················· | ···················· |
| P | 163 hCoV-19/Denmark/DCGC-2359/2020\|EPI_ISL_617699\|2020 | ···················· | ···················· | ····················· | ···················· | ···················· | ···················· |
| P | 164 hCoV-19/Denmark/DCGC-7947/2020\|EPI_ISL_618900\|2020 | ···················· | ···················· | ····················· | ···················· | ···················· | ···················· |
| P | 165 hCoV-19/Denmark/DCGC-7949/2020\|EPI_ISL_618901\|2020 | ···················· | ·A·················· | ····················· | ···················· | ···················· | ··A················· |
| P | 166 hCoV-19/Denmark/DCGC-7953/2020\|EPI_ISL_618902\|2020 | ···················· | ···················· | ····················· | ···················· | ···················· | ···················· |
| P | 167 hCoV-19/Denmark/DCGC-7956/2020\|EPI_ISL_618903\|2020 | ···················· | ···················· | ····················· | ···················· | ···················· | ···················· |
| P | 168 hCoV-19/Denmark/DCGC-7960/2020\|EPI_ISL_618904\|2020 | ···················· | ···················· | ····················· | ···················· | ···················· | ···················· |
| P | 169 hCoV-19/Denmark/DCGC-7316/2020\|EPI_ISL_618957\|2020 | ···················· | ···················· | ····················· | ···················· | ···················· | ···················· |
| P | 170 hCoV-19/Denmark/DCGC-7116/2020\|EPI_ISL_618958\|2020 | ···················· | ···················· | ····················· | ···················· | ···················· | ···················· |
| P | 171 hCoV-19/Denmark/DCGC-7234/2020\|EPI_ISL_618959\|2020 | ···················· | ···················· | ····················· | ···················· | ···················· | ···················· |
| P | 172 hCoV-19/Denmark/DCGC-7480/2020\|EPI_ISL_618960\|2020 | ···················· | ···················· | ····················· | ···················· | ···················· | ···················· |
| P | 173 hCoV-19/Denmark/DCGC-7202/2020\|EPI_ISL_618961\|2020 | ···················· | ···················· | ····················· | ···················· | ···················· | ···················· |
| P | 174 hCoV-19/Denmark/DCGC-7190/2020\|EPI_ISL_618975\|2020 | ···················· | ···················· | ····················· | ···················· | ···················· | ···················· |
| P | 175 hCoV-19/Denmark/DCGC-7230/2020\|EPI_ISL_618976\|2020 | ···················· | ···················· | ····················· | ···················· | ···················· | ···················· |
| P | 176 hCoV-19/Denmark/DCGC-7242/2020\|EPI_ISL_618977\|2020 | ···················· | ···················· | ····················· | ···················· | ···················· | ···················· |
| P | 177 hCoV-19/Denmark/DCGC-1615/2020\|EPI_ISL_618978\|2020 | ···················· | ···················· | ····················· | ···················· | ···················· | ···················· |
| P | 178 hCoV-19/Denmark/DCGC-2049/2020\|EPI_ISL_618979\|2020 | ···················· | ···················· | ····················· | ···················· | ···················· | ···················· |
| P | 179 hCoV-19/Denmark/DCGC-4247/2020\|EPI_ISL_622140\|2020 | ···················· | ···················· | ····················· | ···················· | ···················· | ···················· |
| P | 180 hCoV-19/Denmark/DCGC-4248/2020\|EPI_ISL_622141\|2020 | ···················· | ···················· | ····················· | ···················· | ···················· | ···················· |
| P | 181 hCoV-19/Denmark/DCGC-4249/2020\|EPI_ISL_622142\|2020 | ···················· | ···················· | ····················· | ···················· | ···················· | ···················· |
| P | 182 hCoV-19/Denmark/DCGC-4250/2020\|EPI_ISL_622143\|2020 | ···················· | ···················· | ····················· | ···················· | ···················· | ···················· |
| P | 183 hCoV-19/Denmark/DCGC-4251/2020\|EPI_ISL_622144\|2020 | ···················· | ···················· | ····················· | ···················· | ···················· | ···················· |
| P | 184 hCoV-19/Denmark/DCGC-4252/2020\|EPI_ISL_622145\|2020 | ···················· | ···················· | ····················· | ···················· | ···················· | ···················· |
| P | 185 hCoV-19/Denmark/DCGC-4253/2020\|EPI_ISL_622146\|2020 | ···················· | ···················· | ····················· | ···················· | ···················· | ···················· |
| P | 186 hCoV-19/Denmark/DCGC-4254/2020\|EPI_ISL_622147\|2020 | ···················· | ···················· | ····················· | ···················· | ···················· | ···················· |
| P | 187 hCoV-19/Denmark/DCGC-4255/2020\|EPI_ISL_622148\|2020 | ···················· | ···················· | ····················· | ···················· | ···················· | ···················· |
| P | 188 hCoV-19/Denmark/DCGC-4256/2020\|EPI_ISL_622149\|2020 | ···················· | ···················· | ····················· | ···················· | ···················· | ···················· |
| P | 189 hCoV-19/Denmark/DCGC-4711/2020\|EPI_ISL_622150\|2020 | ···················· | ···················· | ····················· | ···················· | ···················· | ···················· |
| P | 190 hCoV-19/Denmark/DCGC-4712/2020\|EPI_ISL_622151\|2020 | ···················· | ···················· | ····················· | ···················· | ···················· | ···················· |
| P | 191 hCoV-19/Denmark/DCGC-4713/2020\|EPI_ISL_622152\|2020 | ···················· | ···················· | ····················· | ···················· | ···················· | ···················· |
| P | 192 hCoV-19/Denmark/DCGC-4714/2020\|EPI_ISL_622153\|2020 | ···················· | ···················· | ····················· | ···················· | ···················· | ···················· |
| P | 193 hCoV-19/Denmark/DCGC-4715/2020\|EPI_ISL_622154\|2020 | ···················· | ···················· | ····················· | ···················· | ···················· | ···················· |

| | | | | | | | | |
|---|---|---|---|---|---|---|---|---|
| P | 194 | hCoV-19/Denmark/DCGC-4716/2020\|EPI_ISL_622155\|2020 | ···················· | ···················· | ···················· | ···················· | ···················· | ···················· |
| P | 195 | hCoV-19/Denmark/DCGC-4717/2020\|EPI_ISL_622156\|2020 | ···················· | ···················· | ···················· | ···················· | ···················· | ···················· |
| P | 196 | hCoV-19/Denmark/DCGC-4718/2020\|EPI_ISL_622157\|2020 | ···················· | ···················· | ···················· | ···················· | ···················· | ···················· |
| P | 197 | hCoV-19/Denmark/DCGC-4719/2020\|EPI_ISL_622158\|2020 | ···················· | ···················· | ···················· | ···················· | ···················· | ···················· |
| P | 198 | hCoV-19/Denmark/DCGC-4720/2020\|EPI_ISL_622159\|2020 | ···················· | ···················· | ···················· | ···················· | ···················· | ···················· |
| P | 199 | hCoV-19/Denmark/DCGC-4722/2020\|EPI_ISL_622161\|2020 | ···················· | ···················· | ···················· | ···················· | ···················· | ···················· |
| P | 200 | hCoV-19/Denmark/DCGC-4723/2020\|EPI_ISL_622162\|2020 | ···················· | ···················· | ···················· | ···················· | ···················· | ···················· |
| P | 201 | hCoV-19/Denmark/DCGC-4724/2020\|EPI_ISL_622163\|2020 | ···················· | ···················· | ···················· | ···················· | ···················· | ···················· |
| P | 202 | hCoV-19/Denmark/DCGC-4725/2020\|EPI_ISL_622164\|2020 | ···················· | ···················· | ···················· | ···················· | ···················· | ···················· |
| P | 203 | hCoV-19/Denmark/DCGC-4726/2020\|EPI_ISL_622165\|2020 | ···················· | ···················· | ···················· | ···················· | ···················· | ···················· |
| P | 204 | hCoV-19/Denmark/DCGC-4727/2020\|EPI_ISL_622166\|2020 | ···················· | ···················· | ···················· | ···················· | ···················· | ···················· |
| P | 205 | hCoV-19/Denmark/DCGC-4728/2020\|EPI_ISL_622167\|2020 | ···················· | ···················· | ···················· | ···················· | ···················· | ···················· |
| P | 206 | hCoV-19/Denmark/DCGC-4729/2020\|EPI_ISL_622168\|2020 | ···················· | ·A·················· | ···················· | ···················· | ···················· | ···················· |
| P | 207 | hCoV-19/Denmark/DCGC-4730/2020\|EPI_ISL_622169\|2020 | ···················· | ···················· | ···················· | ···················· | ···················· | ···················· |
| P | 208 | hCoV-19/Denmark/DCGC-4731/2020\|EPI_ISL_622170\|2020 | ···················· | ···················· | ···················· | ···················· | ···················· | ···················· |
| P | 209 | hCoV-19/Denmark/DCGC-4732/2020\|EPI_ISL_622171\|2020 | ···················· | ···················· | ···················· | ···················· | ···················· | ···················· |
| P | 210 | hCoV-19/Denmark/DCGC-4733/2020\|EPI_ISL_622172\|2020 | ···················· | ···················· | ···················· | ···················· | ···················· | ···················· |
| P | 211 | hCoV-19/Denmark/DCGC-4734/2020\|EPI_ISL_622173\|2020 | ···················· | ···················· | ···················· | ···················· | ···················· | ···················· |
| P | 212 | hCoV-19/Denmark/DCGC-4735/2020\|EPI_ISL_622174\|2020 | ···················· | ···················· | ···················· | ···················· | ···················· | ···················· |
| P | 213 | hCoV-19/Denmark/DCGC-4738/2020\|EPI_ISL_622177\|2020 | ···················· | ···················· | ···················· | ···················· | ···················· | ···················· |
| P | 214 | hCoV-19/Denmark/DCGC-4739/2020\|EPI_ISL_622178\|2020 | ···················· | ···················· | ···················· | ···················· | ···················· | ···················· |
| P | 215 | hCoV-19/Denmark/DCGC-4740/2020\|EPI_ISL_622179\|2020 | ···················· | ···················· | ···················· | ···················· | ···················· | ···················· |
| P | 216 | hCoV-19/Denmark/DCGC-4741/2020\|EPI_ISL_622180\|2020 | ···················· | ···················· | ···················· | ···················· | ···················· | ···················· |
| P | 217 | hCoV-19/Denmark/DCGC-4742/2020\|EPI_ISL_622181\|2020 | ···················· | ···················· | ···················· | ···················· | ···················· | ···················· |
| P | 218 | hCoV-19/Denmark/DCGC-4743/2020\|EPI_ISL_622182\|2020 | ···················· | ···················· | ···················· | ···················· | ···················· | ···················· |
| P | 219 | hCoV-19/Denmark/DCGC-4744/2020\|EPI_ISL_622183\|2020 | ···················· | ···················· | ···················· | ···················· | ···················· | ···················· |
| P | 220 | hCoV-19/Denmark/DCGC-4746/2020\|EPI_ISL_622185\|2020 | ···················· | ···················· | ···················· | ···················· | ···················· | ···················· |
| P | 221 | hCoV-19/Denmark/DCGC-4747/2020\|EPI_ISL_622186\|2020 | ···················· | ···················· | ···················· | ···················· | ···················· | ···················· |
| P | 222 | hCoV-19/Denmark/DCGC-4748/2020\|EPI_ISL_622187\|2020 | ···················· | ···················· | ···················· | ···················· | ···················· | ···················· |
| P | 223 | hCoV-19/Denmark/DCGC-4749/2020\|EPI_ISL_622188\|2020 | ···················· | ···················· | ···················· | ···················· | ···················· | ···················· |
| P | 224 | hCoV-19/Denmark/DCGC-4750/2020\|EPI_ISL_622189\|2020 | ···················· | ···················· | ···················· | ···················· | ···················· | ···················· |
| P | 225 | hCoV-19/Denmark/DCGC-4751/2020\|EPI_ISL_622190\|2020 | ···················· | ···················· | ···················· | ···················· | ···················· | ···················· |
| P | 226 | hCoV-19/Denmark/DCGC-4752/2020\|EPI_ISL_622191\|2020 | ···················· | ···················· | ···················· | ···················· | ···················· | ···················· |
| P | 227 | hCoV-19/Denmark/DCGC-4753/2020\|EPI_ISL_622192\|2020 | ···················· | ···················· | ···················· | ···················· | ···················· | ···················· |
| P | 228 | hCoV-19/Denmark/DCGC-4754/2020\|EPI_ISL_622193\|2020 | ···················· | ···················· | ···················· | ···················· | ···················· | ···················· |
| P | 229 | hCoV-19/Denmark/DCGC-4755/2020\|EPI_ISL_622194\|2020 | ···················· | ···················· | ···················· | ···················· | ···················· | ···················· |
| P | 230 | hCoV-19/Denmark/DCGC-4757/2020\|EPI_ISL_622196\|2020 | ···················· | ···················· | ···················· | ···················· | ···················· | ···················· |
| P | 231 | hCoV-19/Denmark/DCGC-4758/2020\|EPI_ISL_622197\|2020 | ···················· | ···················· | ···················· | ···················· | ···················· | ···················· |
| P | 232 | hCoV-19/Denmark/DCGC-4759/2020\|EPI_ISL_622198\|2020 | ···················· | ···················· | ···················· | ···················· | ···················· | ···················· |
| P | 233 | hCoV-19/Denmark/DCGC-4760/2020\|EPI_ISL_622199\|2020 | ···················· | ···················· | ···················· | ···················· | ···················· | ···················· |
| P | 234 | hCoV-19/England/MILK-AB743F/2020\|EPI_ISL_623470\|20 | ···················· | ···················· | ···················· | ···················· | ···················· | ···················· |
| P | 235 | hCoV-19/England/MILK-ABB608/2020\|EPI_ISL_623471\|20 | ···················· | ···················· | ···················· | ···················· | ···················· | ···················· |
| P | 236 | hCoV-19/England/MILK-ABB714/2020\|EPI_ISL_623472\|20 | ···················· | ···················· | ···················· | ···················· | ···················· | ···················· |
| P | 237 | hCoV-19/England/MILK-AB75C3/2020\|EPI_ISL_623473\|20 | ···················· | ···················· | ···················· | ···················· | ···················· | ···················· |
| P | 238 | hCoV-19/England/CAMC-AB031B/2020\|EPI_ISL_623474\|20 | ···················· | ···················· | ···················· | ···················· | ···················· | ···················· |
| P | 239 | hCoV-19/England/MILK-AB76EE/2020\|EPI_ISL_623478\|20 | ···················· | ···················· | ···················· | ···················· | ···················· | ···················· |
| P | 240 | hCoV-19/England/MILK-AB8058/2020\|EPI_ISL_623479\|20 | ···················· | ···················· | ···················· | ···················· | ···················· | ···················· |
| P | 241 | hCoV-19/England/MILK-ABB44A/2020\|EPI_ISL_623480\|20 | ···················· | ···················· | ···················· | ···················· | ···················· | ···················· |
| P | 242 | hCoV-19/England/MILK-ABB3D4/2020\|EPI_ISL_623481\|20 | ···················· | ···················· | ···················· | ···················· | ······T············· | ···················· |
| P | 243 | hCoV-19/England/CAMC-AAFEC3/2020\|EPI_ISL_623482\|20 | ···················· | ···················· | ···················· | ···················· | ···················· | ···················· |
| P | 244 | hCoV-19/England/MILK-AB8261/2020\|EPI_ISL_623483\|20 | ···················· | ···················· | ···················· | ···················· | ···················· | ···················· |
| P | 245 | hCoV-19/England/CAMC-AAFEE1/2020\|EPI_ISL_623485\|20 | ···················· | ···················· | ···················· | ···················· | ···················· | ···················· |
| P | 246 | hCoV-19/England/MILK-AB7824/2020\|EPI_ISL_623487\|20 | ···················· | ·A·················· | ···················· | ···················· | ···················· | ···················· |
| P | 247 | hCoV-19/England/CAMC-AAFD6C/2020\|EPI_ISL_623488\|20 | ···················· | ···················· | ···················· | ···················· | ······T············· | ···················· |
| P | 248 | hCoV-19/England/CAMC-AB0357/2020\|EPI_ISL_623489\|20 | ···················· | ···················· | ···················· | ···················· | ···················· | ···················· |
| P | 249 | hCoV-19/England/CAMC-AAFAFC/2020\|EPI_ISL_623490\|20 | ···················· | ···················· | ···K················ | ···················· | ···················· | ···················· |
| P | 250 | hCoV-19/England/MILK-AB84B6/2020\|EPI_ISL_623491\|20 | ···················· | ···················· | ···················· | ···················· | ···················· | ···················· |
| P | 251 | hCoV-19/England/MILK-AB77EB/2020\|EPI_ISL_623492\|20 | ···················· | ···················· | ···················· | ···················· | ···················· | ···················· |
| P | 252 | hCoV-19/England/MILK-ABB2C8/2020\|EPI_ISL_623493\|20 | ···················· | ···················· | ···················· | ···················· | ···················· | ···················· |
| P | 253 | hCoV-19/England/CAMC-AAFE78/2020\|EPI_ISL_623494\|20 | ···················· | ···················· | ···················· | ···················· | ···················· | ···················· |
| P | 254 | hCoV-19/England/CAMC-AAFB44/2020\|EPI_ISL_623495\|20 | ···················· | ···················· | ···················· | ···················· | ···················· | ···················· |
| P | 255 | hCoV-19/England/MILK-AB76DF/2020\|EPI_ISL_623496\|20 | ···················· | ···················· | ···················· | ···················· | ······T············· | ···················· |
| P | 256 | hCoV-19/England/MILK-AB7772/2020\|EPI_ISL_623499\|20 | ···················· | ···················· | ···················· | ···················· | ······T············· | ···················· |
| P | 257 | hCoV-19/Gibraltar/204462825/2020\|EPI_ISL_615000\|20 | ···················· | ···················· | ···················· | ···················· | ···················· | ···················· |
| P | 258 | hCoV-19/Gibraltar/204462826/2020\|EPI_ISL_615001\|20 | ···················· | ···················· | ···················· | ···················· | ···················· | ···················· |
| P | 259 | hCoV-19/Gibraltar/204462827/2020\|EPI_ISL_615002\|20 | ···················· | ···················· | ···················· | ···················· | ···················· | ···················· |
| P | 260 | hCoV-19/Gibraltar/204462828/2020\|EPI_ISL_615003\|20 | ···················· | ···················· | ···················· | ···················· | ···················· | ···················· |
| P | 261 | hCoV-19/Gibraltar/204462830/2020\|EPI_ISL_615005\|20 | ···················· | ···················· | ···················· | ···················· | ···················· | ···················· |
| P | 262 | hCoV-19/Gibraltar/204462831/2020\|EPI_ISL_615006\|20 | ···················· | ···················· | ···················· | ···················· | ···················· | ···················· |
| P | 263 | hCoV-19/Gibraltar/204462833/2020\|EPI_ISL_615007\|20 | ···················· | ···················· | ···················· | ···················· | ···················· | ···················· |
| P | 264 | hCoV-19/Gibraltar/204462834/2020\|EPI_ISL_615008\|20 | ···················· | ···················· | ···················· | ···················· | ···················· | ···················· |
| P | 265 | hCoV-19/Gibraltar/204462835/2020\|EPI_ISL_615009\|20 | ···················· | ···················· | ···················· | ···················· | ···················· | ···················· |
| P | 266 | hCoV-19/Denmark/DCGC-3473/2020\|EPI_ISL_616260\|2020 | ···················· | ·A·················· | ···················· | ···················· | ···················· | ···A················ |
| P | 267 | hCoV-19/Denmark/DCGC-3474/2020\|EPI_ISL_616261\|2020 | ···················· | ···················· | ···················· | ···················· | ···················· | ···················· |
| P | 268 | hCoV-19/Denmark/DCGC-3475/2020\|EPI_ISL_616262\|2020 | ···················· | ···················· | ···················· | ···················· | ···················· | ···················· |
| P | 269 | hCoV-19/Denmark/DCGC-3476/2020\|EPI_ISL_616263\|2020 | ···················· | ···················· | ···················· | ···················· | ···················· | ···················· |
| P | 270 | hCoV-19/Denmark/DCGC-3477/2020\|EPI_ISL_616264\|2020 | ···················· | ···················· | ···················· | ···················· | ···················· | ···················· |
| P | 271 | hCoV-19/Denmark/DCGC-3269/2020\|EPI_ISL_616265\|2020 | ···················· | ···················· | ···················· | ···················· | ···················· | ···················· |
| P | 272 | hCoV-19/Denmark/DCGC-3478/2020\|EPI_ISL_616266\|2020 | ···················· | ···················· | ···················· | ···················· | ···················· | ···················· |
| P | 273 | hCoV-19/Denmark/DCGC-3479/2020\|EPI_ISL_616267\|2020 | ···················· | ···················· | ···················· | ···················· | ···················· | ···················· |

| | | | | | | | |
|---|---|---|---|---|---|---|---|
| P | 274 hCoV-19/Denmark/DCGC-3480/2020\|EPI_ISL_616268\|2020 | ···················· | ···················· | ···················· | ···················· | ···················· | ···················· |
| P | 275 hCoV-19/Denmark/DCGC-3482/2020\|EPI_ISL_616270\|2020 | ···················· | ···················· | ···················· | ···················· | ···················· | ···················· |
| P | 276 hCoV-19/Denmark/DCGC-3483/2020\|EPI_ISL_616271\|2020 | ···················· | ···················· | ···················· | ···················· | ···················· | ···················· |
| P | 277 hCoV-19/Denmark/DCGC-3484/2020\|EPI_ISL_616272\|2020 | ···················· | ···················· | ···················· | ···················· | ···················· | ···················· |
| P | 278 hCoV-19/Denmark/DCGC-3485/2020\|EPI_ISL_616273\|2020 | ···················· | ···G················ | ···················· | ···················· | ···················· | ···················· |
| P | 279 hCoV-19/Denmark/DCGC-3486/2020\|EPI_ISL_616274\|2020 | ···················· | ···················· | ···················· | ···················· | ···················· | ···················· |
| P | 280 hCoV-19/Denmark/DCGC-3487/2020\|EPI_ISL_616275\|2020 | ···················· | ···················· | ···················· | ···················· | ···················· | ···················· |
| P | 281 hCoV-19/Denmark/DCGC-3488/2020\|EPI_ISL_616276\|2020 | ···················· | ···················· | ···················· | ···················· | ···················· | ···················· |
| P | 282 hCoV-19/Denmark/DCGC-3489/2020\|EPI_ISL_616277\|2020 | ···················· | ···················· | ···················· | ···················· | ···················· | ···················· |
| P | 283 hCoV-19/Denmark/DCGC-3490/2020\|EPI_ISL_616278\|2020 | ···················· | ···················· | ···················· | ···················· | ···················· | ···················· |
| P | 284 hCoV-19/Denmark/DCGC-3491/2020\|EPI_ISL_616279\|2020 | ···················· | ···················· | ···················· | ···················· | ···················· | ···················· |
| P | 285 hCoV-19/Denmark/DCGC-3492/2020\|EPI_ISL_616280\|2020 | ···················· | ·A·················· | ···················· | ···················· | ···················· | ··A················· |
| P | 286 hCoV-19/Denmark/DCGC-3494/2020\|EPI_ISL_616282\|2020 | ···················· | ···················· | ···················· | ···················· | ···················· | ···················· |
| P | 287 hCoV-19/Denmark/DCGC-3495/2020\|EPI_ISL_616283\|2020 | ···················· | ···················· | ···················· | ···················· | ···················· | ···················· |
| P | 288 hCoV-19/Denmark/DCGC-3496/2020\|EPI_ISL_616284\|2020 | ···················· | ···················· | ···················· | ···················· | ···················· | ···················· |
| P | 289 hCoV-19/Denmark/DCGC-3270/2020\|EPI_ISL_616285\|2020 | ···················· | ···················· | ···················· | ···················· | ···················· | ···················· |
| P | 290 hCoV-19/Denmark/DCGC-3497/2020\|EPI_ISL_616286\|2020 | ···················· | ···················· | ···················· | ···················· | ···················· | ···················· |
| P | 291 hCoV-19/Denmark/DCGC-3498/2020\|EPI_ISL_616287\|2020 | ···················· | ···················· | ···················· | ···················· | ···················· | ···················· |
| P | 292 hCoV-19/Denmark/DCGC-3499/2020\|EPI_ISL_616288\|2020 | ···················· | ···················· | ···················· | ···················· | ···················· | ···················· |
| P | 293 hCoV-19/Denmark/DCGC-3500/2020\|EPI_ISL_616289\|2020 | ···················· | ···················· | ···················· | ···················· | ···················· | ···················· |
| P | 294 hCoV-19/Denmark/DCGC-3501/2020\|EPI_ISL_616290\|2020 | ···················· | ···················· | ···················· | ···················· | ···················· | ···················· |
| P | 295 hCoV-19/Denmark/DCGC-3502/2020\|EPI_ISL_616291\|2020 | ···················· | ···················· | ···················· | ···················· | ···················· | ···················· |
| P | 296 hCoV-19/Denmark/DCGC-3504/2020\|EPI_ISL_616293\|2020 | ···················· | ···················· | ···················· | ···················· | ···················· | ···················· |
| P | 297 hCoV-19/Denmark/DCGC-3505/2020\|EPI_ISL_616294\|2020 | ···················· | ···················· | ···················· | ···················· | ···················· | ···················· |
| P | 298 hCoV-19/Denmark/DCGC-3507/2020\|EPI_ISL_616296\|2020 | ···················· | ···················· | ···················· | ···················· | ···················· | ···················· |
| P | 299 hCoV-19/Denmark/DCGC-3508/2020\|EPI_ISL_616297\|2020 | ···················· | ·A·················· | ···················· | ···················· | ···················· | ···················· |
| P | 300 hCoV-19/Denmark/DCGC-3509/2020\|EPI_ISL_616298\|2020 | ···················· | ···················· | ···················· | ···················· | ···················· | ···················· |
| P | 301 hCoV-19/Denmark/DCGC-3510/2020\|EPI_ISL_616299\|2020 | ···················· | ···················· | ···················· | ···················· | ···················· | ···················· |
| P | 302 hCoV-19/Denmark/DCGC-3519/2020\|EPI_ISL_616310\|2020 | ···················· | ···················· | ···················· | ···················· | ···················· | ···················· |
| P | 303 hCoV-19/Denmark/DCGC-3520/2020\|EPI_ISL_616311\|2020 | ···················· | ···················· | ···················· | ···················· | ···················· | ···················· |
| P | 304 hCoV-19/Denmark/DCGC-3521/2020\|EPI_ISL_616312\|2020 | ···················· | ···················· | ···················· | ···················· | ···················· | ···················· |
| P | 305 hCoV-19/Denmark/DCGC-3529/2020\|EPI_ISL_616320\|2020 | ···················· | ···················· | ···················· | ···················· | ···················· | ···················· |
| P | 306 hCoV-19/Denmark/DCGC-3530/2020\|EPI_ISL_616321\|2020 | ···················· | ···················· | ···················· | ···················· | ···················· | ···················· |
| P | 307 hCoV-19/Denmark/DCGC-3531/2020\|EPI_ISL_616322\|2020 | ···················· | ···················· | ···················· | ···················· | ···················· | ···················· |
| P | 308 hCoV-19/Denmark/DCGC-3532/2020\|EPI_ISL_616323\|2020 | ···················· | ···················· | ···················· | ···················· | ···················· | ···················· |
| P | 309 hCoV-19/Denmark/DCGC-3273/2020\|EPI_ISL_616324\|2020 | ···················· | ···················· | ···················· | ···················· | ···················· | ···················· |
| P | 310 hCoV-19/Denmark/DCGC-3533/2020\|EPI_ISL_616325\|2020 | ···················· | ·A·················· | ···················· | ···················· | ···················· | ··A················· |
| P | 311 hCoV-19/Denmark/DCGC-3274/2020\|EPI_ISL_616326\|2020 | ···················· | ···················· | ···················· | ···················· | ···················· | ···················· |
| P | 312 hCoV-19/Denmark/DCGC-3534/2020\|EPI_ISL_616327\|2020 | ···················· | ···················· | ···················· | ···················· | ···················· | ···················· |
| P | 313 hCoV-19/Denmark/DCGC-3535/2020\|EPI_ISL_616328\|2020 | ···················· | ···················· | ···················· | ···················· | ···················· | ···················· |
| P | 314 hCoV-19/Denmark/DCGC-3536/2020\|EPI_ISL_616329\|2020 | ···················· | ···················· | ···················· | ···················· | ···················· | ···················· |
| P | 315 hCoV-19/Denmark/DCGC-3537/2020\|EPI_ISL_616330\|2020 | ···················· | ···················· | ···················· | ···················· | ···················· | ···················· |
| P | 316 hCoV-19/Denmark/DCGC-3538/2020\|EPI_ISL_616331\|2020 | ···················· | ···················· | ···················· | ···················· | ···················· | ···················· |
| P | 317 hCoV-19/Denmark/DCGC-3539/2020\|EPI_ISL_616332\|2020 | ···················· | ···················· | ···················· | ···················· | ···················· | ···················· |
| P | 318 hCoV-19/Denmark/DCGC-3540/2020\|EPI_ISL_616333\|2020 | ···················· | ···················· | ···················· | ···················· | ···················· | ···················· |
| P | 319 hCoV-19/Denmark/DCGC-3541/2020\|EPI_ISL_616334\|2020 | ···················· | ···················· | ···················· | ···················· | ···················· | ···················· |
| P | 320 hCoV-19/Denmark/DCGC-3543/2020\|EPI_ISL_616336\|2020 | ···················· | ···················· | ···················· | ···················· | ···················· | ···················· |
| P | 321 hCoV-19/Denmark/DCGC-3544/2020\|EPI_ISL_616337\|2020 | ···················· | ···················· | ···················· | ···················· | ···················· | ···················· |
| P | 322 hCoV-19/Denmark/DCGC-3545/2020\|EPI_ISL_616338\|2020 | ···········T········ | ···················· | ···················· | ···················· | ···················· | ···················· |
| P | 323 hCoV-19/Denmark/DCGC-3546/2020\|EPI_ISL_616339\|2020 | ···················· | ···················· | ···················· | ···················· | ···················· | ···················· |
| P | 324 hCoV-19/Denmark/DCGC-2408/2020\|EPI_ISL_617590\|2020 | ···················· | ···················· | ···················· | ···················· | ···················· | ···················· |
| P | 325 hCoV-19/Denmark/DCGC-2409/2020\|EPI_ISL_617591\|2020 | ···················· | ···················· | ···················· | ···················· | ···················· | ···················· |
| P | 326 hCoV-19/Denmark/DCGC-2410/2020\|EPI_ISL_617592\|2020 | ···················· | ···················· | ···················· | ···················· | ···················· | ···················· |
| P | 327 hCoV-19/Denmark/DCGC-2411/2020\|EPI_ISL_617593\|2020 | ···················· | ···················· | ···················· | ···················· | ···················· | ···················· |
| P | 328 hCoV-19/Denmark/DCGC-2412/2020\|EPI_ISL_617594\|2020 | ···················· | ···················· | ···················· | ···················· | ···················· | ···················· |
| P | 329 hCoV-19/Denmark/DCGC-2413/2020\|EPI_ISL_617595\|2020 | ···················· | ···················· | ···················· | ···················· | ···················· | ···················· |
| P | 330 hCoV-19/Denmark/DCGC-2414/2020\|EPI_ISL_617596\|2020 | ···················· | ···················· | ···················· | ···················· | ···················· | ···················· |
| P | 331 hCoV-19/Denmark/DCGC-2415/2020\|EPI_ISL_617597\|2020 | ···················· | ···················· | ···················· | ···················· | ···················· | ···················· |
| P | 332 hCoV-19/Denmark/DCGC-2416/2020\|EPI_ISL_617598\|2020 | ···················· | ···················· | ···················· | ···················· | ···················· | ···················· |
| P | 333 hCoV-19/Denmark/DCGC-2283/2020\|EPI_ISL_617641\|2020 | ···················· | ···················· | ···················· | ···················· | ···················· | ···················· |
| P | 334 hCoV-19/Denmark/DCGC-2284/2020\|EPI_ISL_617642\|2020 | ···················· | ···················· | ···················· | ···················· | ···················· | ···················· |
| P | 335 hCoV-19/Denmark/DCGC-2313/2020\|EPI_ISL_617650\|2020 | ···················· | ···················· | ···················· | ···················· | ···················· | ···················· |
| P | 336 hCoV-19/Denmark/DCGC-2314/2020\|EPI_ISL_617651\|2020 | ···················· | ···················· | ···················· | ···················· | ···················· | ···················· |
| P | 337 hCoV-19/Denmark/DCGC-2316/2020\|EPI_ISL_617653\|2020 | ···················· | ···················· | ···················· | ···················· | ···················· | ···················· |
| P | 338 hCoV-19/Denmark/DCGC-2317/2020\|EPI_ISL_617654\|2020 | ···················· | ···················· | ···················· | ···················· | ···················· | ···················· |
| P | 339 hCoV-19/Denmark/DCGC-2319/2020\|EPI_ISL_617656\|2020 | ···················· | ···················· | ···················· | ···················· | ···················· | ···················· |
| P | 340 hCoV-19/Denmark/DCGC-2320/2020\|EPI_ISL_617657\|2020 | ···················· | ···················· | ···················· | ···················· | ···················· | ···················· |
| P | 341 hCoV-19/Denmark/DCGC-2322/2020\|EPI_ISL_617659\|2020 | ···················· | ···················· | ···················· | ···················· | ···················· | ···················· |
| P | 342 hCoV-19/Denmark/DCGC-2324/2020\|EPI_ISL_617661\|2020 | ···················· | ···················· | ···················· | ···················· | ···················· | ···················· |
| P | 343 hCoV-19/Denmark/DCGC-2325/2020\|EPI_ISL_617662\|2020 | ···················· | ···················· | ···················· | ···················· | ···················· | ···················· |
| P | 344 hCoV-19/Denmark/DCGC-2326/2020\|EPI_ISL_617663\|2020 | ···················· | ···················· | ···················· | ···················· | ···················· | ···················· |
| P | 345 hCoV-19/Denmark/DCGC-2327/2020\|EPI_ISL_617664\|2020 | ···················· | ···················· | ···················· | ···················· | ···················· | ···················· |
| P | 346 hCoV-19/Denmark/DCGC-2328/2020\|EPI_ISL_617665\|2020 | ···················· | ···················· | ···················· | ···················· | ···················· | ···················· |
| P | 347 hCoV-19/Denmark/DCGC-2329/2020\|EPI_ISL_617666\|2020 | ···················· | ···················· | ···················· | ···················· | ···················· | ···················· |
| P | 348 hCoV-19/Denmark/DCGC-2330/2020\|EPI_ISL_617667\|2020 | ···················· | ···················· | ···················· | ···················· | ···················· | ···················· |
| P | 349 hCoV-19/Denmark/DCGC-2331/2020\|EPI_ISL_617668\|2020 | ···················· | ···················· | ···················· | ···················· | ···················· | ···················· |
| P | 350 hCoV-19/Denmark/DCGC-2689/2020\|EPI_ISL_617669\|2020 | ···················· | ···················· | ···················· | ···················· | ···················· | ···················· |
| P | 351 hCoV-19/Denmark/DCGC-7104/2020\|EPI_ISL_618974\|2020 | ···················· | ···················· | ···················· | ···················· | ···················· | ···················· |
| P | 352 hCoV-19/Denmark/DCGC-2050/2020\|EPI_ISL_618980\|2020 | ···················· | ···················· | ···················· | ···················· | ···················· | ···················· |
| P | 353 hCoV-19/Denmark/DCGC-7069/2020\|EPI_ISL_618981\|2020 | ···················· | ···················· | ···················· | ···················· | ···················· | ···················· |

| | | | | | | | |
|---|---|---|---|---|---|---|---|
| P | 354 hCoV-19/Denmark/DCGC-7082/2020\|EPI_ISL_618982\|2020 | ···················· | ···················· | ···················· | ···················· | ···················· | ···················· |
| P | 355 hCoV-19/Denmark/DCGC-7086/2020\|EPI_ISL_618983\|2020 | ···················· | ···················· | ···················· | ···················· | ···················· | ···················· |
| P | 356 hCoV-19/Denmark/DCGC-7099/2020\|EPI_ISL_618984\|2020 | ···················· | ···················· | ···················· | ···················· | ···················· | ···················· |
| P | 357 hCoV-19/Denmark/DCGC-7106/2020\|EPI_ISL_618985\|2020 | ···················· | ···················· | ···················· | ···················· | ···················· | ···················· |
| P | 358 hCoV-19/Denmark/DCGC-7109/2020\|EPI_ISL_618986\|2020 | ···················· | ···················· | ···················· | ···················· | ···················· | ···················· |
| P | 359 hCoV-19/Denmark/DCGC-7117/2020\|EPI_ISL_618987\|2020 | ···················· | ···················· | ···················· | ···················· | ···················· | ···················· |
| P | 360 hCoV-19/Denmark/DCGC-7134/2020\|EPI_ISL_618988\|2020 | ···················· | ···················· | ···················· | ···················· | ···················· | ···················· |
| P | 361 hCoV-19/Denmark/DCGC-7135/2020\|EPI_ISL_618989\|2020 | ···················· | ···················· | ···················· | ···················· | ···················· | ···················· |
| P | 362 hCoV-19/Denmark/DCGC-7137/2020\|EPI_ISL_618990\|2020 | ···················· | ···················· | ···················· | ···················· | ···················· | ···················· |
| P | 363 hCoV-19/Denmark/DCGC-7143/2020\|EPI_ISL_618991\|2020 | ···················· | ···················· | ···················· | ···················· | ···················· | ···················· |
| P | 364 hCoV-19/Denmark/DCGC-7150/2020\|EPI_ISL_618992\|2020 | ···················· | ···················· | ···················· | ···················· | ···················· | ···················· |
| P | 365 hCoV-19/Denmark/DCGC-7164/2020\|EPI_ISL_618993\|2020 | ···················· | ···················· | ···················· | ···················· | ···················· | ···················· |
| P | 366 hCoV-19/Denmark/DCGC-7167/2020\|EPI_ISL_618994\|2020 | ···················· | ···················· | ···················· | ···················· | ···················· | ···················· |
| P | 367 hCoV-19/Denmark/DCGC-7169/2020\|EPI_ISL_618995\|2020 | ···················· | ···················· | ···················· | ···················· | ···················· | ···················· |
| P | 368 hCoV-19/Denmark/DCGC-7173/2020\|EPI_ISL_618996\|2020 | ···················· | ···················· | ···················· | ···················· | ···················· | ···················· |
| P | 369 hCoV-19/Denmark/DCGC-7174/2020\|EPI_ISL_618997\|2020 | ···················· | ···················· | ···················· | ···················· | ···················· | ···················· |
| P | 370 hCoV-19/Denmark/DCGC-7178/2020\|EPI_ISL_618998\|2020 | ···················· | ···················· | ···················· | ···················· | ···················· | ···················· |
| P | 371 hCoV-19/Denmark/DCGC-7192/2020\|EPI_ISL_618999\|2020 | ···················· | ···················· | ···················· | ···················· | ···················· | ···················· |
| P | 372 hCoV-19/Denmark/DCGC-4207/2020\|EPI_ISL_622100\|2020 | ···················· | ···················· | ···················· | ···················· | ···················· | ···················· |
| P | 373 hCoV-19/Denmark/DCGC-4208/2020\|EPI_ISL_622101\|2020 | ···················· | ···················· | ···················· | ···················· | ···················· | ···················· |
| P | 374 hCoV-19/Denmark/DCGC-4210/2020\|EPI_ISL_622103\|2020 | ···················· | ···················· | ···················· | ···················· | ···················· | ···················· |
| P | 375 hCoV-19/Denmark/DCGC-4211/2020\|EPI_ISL_622104\|2020 | ···················· | ···················· | ···················· | ···················· | ···················· | ···················· |
| P | 376 hCoV-19/Denmark/DCGC-4212/2020\|EPI_ISL_622105\|2020 | ···················· | ···················· | ···················· | ···················· | ···················· | ···················· |
| P | 377 hCoV-19/Denmark/DCGC-4213/2020\|EPI_ISL_622106\|2020 | ···················· | ···················· | ···················· | ···················· | ···················· | ···················· |
| P | 378 hCoV-19/Denmark/DCGC-4214/2020\|EPI_ISL_622107\|2020 | ···················· | ···················· | ···················· | ···················· | ···················· | ···················· |
| P | 379 hCoV-19/Denmark/DCGC-4215/2020\|EPI_ISL_622108\|2020 | ···················· | ···················· | ···················· | ···················· | ···················· | ···················· |
| P | 380 hCoV-19/Denmark/DCGC-4216/2020\|EPI_ISL_622109\|2020 | ···················· | ···················· | ···················· | ···················· | ···················· | ···················· |
| P | 381 hCoV-19/Denmark/DCGC-4217/2020\|EPI_ISL_622110\|2020 | ···················· | ···················· | ···················· | ···················· | ···················· | ···················· |
| P | 382 hCoV-19/Denmark/DCGC-4218/2020\|EPI_ISL_622111\|2020 | ···················· | ···················· | ···················· | ···················· | ···················· | ···················· |
| P | 383 hCoV-19/Denmark/DCGC-4219/2020\|EPI_ISL_622112\|2020 | ···················· | ···················· | ···················· | ···················· | ···················· | ···················· |
| P | 384 hCoV-19/Denmark/DCGC-4220/2020\|EPI_ISL_622113\|2020 | ···················· | ···················· | ···················· | ···················· | ···················· | ···················· |
| P | 385 hCoV-19/Denmark/DCGC-4221/2020\|EPI_ISL_622114\|2020 | ···················· | ···················· | ···················· | ···················· | ···················· | ···················· |
| P | 386 hCoV-19/Denmark/DCGC-4222/2020\|EPI_ISL_622115\|2020 | ···················· | ···················· | ···················· | ···················· | ···················· | ···················· |
| P | 387 hCoV-19/Denmark/DCGC-4223/2020\|EPI_ISL_622116\|2020 | ···················· | ···················· | ···················· | ···················· | ···················· | ···················· |
| P | 388 hCoV-19/Denmark/DCGC-4224/2020\|EPI_ISL_622117\|2020 | ···················· | ···················· | ···················· | ···················· | ···················· | ···················· |
| P | 389 hCoV-19/Denmark/DCGC-4225/2020\|EPI_ISL_622118\|2020 | ···················· | ···················· | ···················· | ···················· | ···················· | ···················· |
| P | 390 hCoV-19/Denmark/DCGC-4226/2020\|EPI_ISL_622119\|2020 | ···················· | ···················· | ···················· | ···················· | ···················· | ···················· |
| P | 391 hCoV-19/Denmark/DCGC-4227/2020\|EPI_ISL_622120\|2020 | ···················· | ···················· | ···················· | ···················· | ···················· | ···················· |
| P | 392 hCoV-19/Denmark/DCGC-4228/2020\|EPI_ISL_622121\|2020 | ···················· | ···················· | ···················· | ···················· | ···················· | ···················· |
| P | 393 hCoV-19/Denmark/DCGC-4229/2020\|EPI_ISL_622122\|2020 | ···················· | ···················· | ···················· | ···················· | ···················· | ···················· |
| P | 394 hCoV-19/Denmark/DCGC-4230/2020\|EPI_ISL_622123\|2020 | ···················· | ···················· | ···················· | ···················· | ···················· | ···················· |
| P | 395 hCoV-19/Denmark/DCGC-4231/2020\|EPI_ISL_622124\|2020 | ···················· | ···················· | ···················· | ···················· | ···················· | ···················· |
| P | 396 hCoV-19/Denmark/DCGC-4232/2020\|EPI_ISL_622125\|2020 | ···················· | ···················· | ···················· | ···················· | ···················· | ···················· |
| P | 397 hCoV-19/Denmark/DCGC-4233/2020\|EPI_ISL_622126\|2020 | ···················· | ···················· | ···················· | ···················· | ···················· | ···················· |
| P | 398 hCoV-19/Denmark/DCGC-4234/2020\|EPI_ISL_622127\|2020 | ···················· | ···················· | ···················· | ···················· | ···················· | ···················· |
| P | 399 hCoV-19/Denmark/DCGC-4236/2020\|EPI_ISL_622129\|2020 | ···················· | ···················· | ···················· | ···················· | ···················· | ···················· |
| P | 400 hCoV-19/Denmark/DCGC-4237/2020\|EPI_ISL_622130\|2020 | ···················· | ···················· | ···················· | ···················· | ···················· | ···················· |
| P | 401 hCoV-19/Denmark/DCGC-4239/2020\|EPI_ISL_622132\|2020 | ···················· | ···················· | ···················· | ···················· | ···················· | ···················· |
| P | 402 hCoV-19/Denmark/DCGC-4240/2020\|EPI_ISL_622133\|2020 | ···················· | ···················· | ···················· | ···················· | ···················· | ···················· |
| P | 403 hCoV-19/Denmark/DCGC-4241/2020\|EPI_ISL_622134\|2020 | ···················· | ···················· | ···················· | ···················· | ···················· | ···················· |
| P | 404 hCoV-19/Denmark/DCGC-4242/2020\|EPI_ISL_622135\|2020 | ···················· | ···················· | ···················· | ···················· | ···················· | ···················· |
| P | 405 hCoV-19/Denmark/DCGC-4243/2020\|EPI_ISL_622136\|2020 | ···················· | ···················· | ···················· | ···················· | ···················· | ···················· |
| P | 406 hCoV-19/Denmark/DCGC-4244/2020\|EPI_ISL_622137\|2020 | ···················· | ···················· | ···················· | ···················· | ···················· | ···················· |
| P | 407 hCoV-19/Denmark/DCGC-4245/2020\|EPI_ISL_622138\|2020 | ···················· | ···················· | ···················· | ···················· | ···················· | ···················· |
| P | 408 hCoV-19/Denmark/DCGC-4246/2020\|EPI_ISL_622139\|2020 | ···················· | ···················· | ···················· | ···················· | ···················· | ···················· |
| P | 409 hCoV-19/England/MILK-ABB592/2020\|EPI_ISL_623400\|20 | ···················· | ·A·················· | ···················· | ···················· | ···················· | ··A················· |
| P | 410 hCoV-19/England/MILK-AB8717/2020\|EPI_ISL_623401\|20 | ···················· | ···················· | ···················· | ···················· | ···················· | ···················· |
| P | 411 hCoV-19/England/MILK-AB73D8/2020\|EPI_ISL_623402\|20 | ···················· | ···················· | ···················· | ···················· | ·····T·············· | ···················· |
| P | 412 hCoV-19/England/MILK-AB8216/2020\|EPI_ISL_623403\|20 | ···················· | ···················· | ···················· | ···················· | ···················· | ···················· |
| P | 413 hCoV-19/England/CAMC-AAFD5D/2020\|EPI_ISL_623404\|20 | ···················· | ···················· | ···················· | ···················· | ···················· | ···················· |
| P | 414 hCoV-19/England/CAMC-AB0375/2020\|EPI_ISL_623427\|20 | ···················· | ···················· | ···················· | ···················· | ···················· | ···················· |
| P | 415 hCoV-19/England/MILK-AB7790/2020\|EPI_ISL_623429\|20 | ···················· | ···················· | ···················· | ···················· | ···················· | ···················· |
| P | 416 hCoV-19/England/MILK-ABB811/2020\|EPI_ISL_623430\|20 | ···················· | ···················· | ···················· | ···················· | ···················· | ···················· |
| P | 417 hCoV-19/England/MILK-ABB2E6/2020\|EPI_ISL_623431\|20 | ···················· | ···················· | ···················· | ···················· | ···················· | ···················· |
| P | 418 hCoV-19/England/MILK-AB745D/2020\|EPI_ISL_623432\|20 | ···················· | ···················· | ···················· | ···················· | ···················· | ···················· |
| P | 419 hCoV-19/England/CAMC-AAFB08/2020\|EPI_ISL_623433\|20 | ···················· | ···················· | ···················· | ···················· | ···················· | ···················· |
| P | 420 hCoV-19/England/MILK-AB76A2/2020\|EPI_ISL_623434\|20 | ···················· | ···················· | ···················· | ···················· | ···················· | ···················· |
| P | 421 hCoV-19/England/MILK-AB84A7/2020\|EPI_ISL_623435\|20 | ···················· | ···················· | ···················· | ···················· | ···················· | ···················· |
| P | 422 hCoV-19/England/MILK-AB7E5F/2020\|EPI_ISL_623436\|20 | ···················· | ···················· | ···················· | ···················· | ···················· | ···················· |
| P | 423 hCoV-19/England/CAMC-AB0366/2020\|EPI_ISL_623437\|20 | ···················· | ···················· | ···················· | ···················· | ·····T·············· | ···················· |
| P | 424 hCoV-19/England/MILK-AB81EC/2020\|EPI_ISL_623439\|20 | ···················· | ···················· | ···················· | ···················· | ·····T·············· | ···················· |
| P | 425 hCoV-19/England/CAMC-AAFC9C/2020\|EPI_ISL_623440\|20 | ···················· | ···················· | ···················· | ···················· | ···················· | ···················· |
| P | 426 hCoV-19/England/MILK-AB8119/2020\|EPI_ISL_623442\|20 | ···················· | ···················· | ···················· | ···················· | ···················· | ···················· |
| P | 427 hCoV-19/England/MILK-ABBAA2/2020\|EPI_ISL_623443\|20 | ···················· | ···················· | ···················· | ···················· | ···················· | ···················· |
| P | 428 hCoV-19/England/CAMC-AAFB62/2020\|EPI_ISL_623444\|20 | ···················· | ···················· | ···················· | ···················· | ···················· | ···················· |
| P | 429 hCoV-19/England/CAMC-AAFC05/2020\|EPI_ISL_623445\|20 | ···················· | ···················· | ···················· | ···················· | ···················· | ···················· |
| P | 430 hCoV-19/England/MILK-AB7AD3/2020\|EPI_ISL_623446\|20 | ···················· | ···················· | ···················· | ···················· | ···················· | ···················· |
| P | 431 hCoV-19/England/MILK-AB8410/2020\|EPI_ISL_623447\|20 | ···················· | ···················· | ···················· | ···················· | ···················· | ···················· |
| P | 432 hCoV-19/England/CAMC-AAFDD5/2020\|EPI_ISL_623448\|20 | ···················· | ···················· | ···················· | ···················· | ···················· | ···················· |
| P | 433 hCoV-19/England/CAMC-AAFD02/2020\|EPI_ISL_623449\|20 | ···················· | ···················· | ···················· | ···················· | ·····T·············· | ···················· |

| | | | | | | | | |
|---|---|---|---|---|---|---|---|---|
| P | 434 | hCoV-19/England/CAMC-AAFE1E/2020\|EPI_ISL_623451\|20 | ···················· | ·A·················· | ······················ | ···················· | ···················· | ···A················ |
| P | 435 | hCoV-19/England/MILK-AB77AF/2020\|EPI_ISL_623452\|20 | ···················· | ···················· | ······················ | ···················· | ···················· | ···················· |
| P | 436 | hCoV-19/England/MILK-AB836E/2020\|EPI_ISL_623454\|20 | ···················· | ···················· | ······················ | ···················· | ···················· | ···················· |
| P | 437 | hCoV-19/England/MILK-AB7AE2/2020\|EPI_ISL_623455\|20 | ···················· | ···················· | ······················ | ···················· | ······T············· | ···················· |
| P | 438 | hCoV-19/England/MILK-AB7A79/2020\|EPI_ISL_623457\|20 | ···················· | ···················· | ······················ | ···················· | ···················· | ···················· |
| P | 439 | hCoV-19/England/MILK-ABB969/2020\|EPI_ISL_623459\|20 | ···················· | ···················· | ······················ | ···················· | ···················· | ···················· |
| P | 440 | hCoV-19/England/MILK-ABB680/2020\|EPI_ISL_623460\|20 | ···················· | ···················· | ······················ | ···················· | ···················· | ···················· |
| P | 441 | hCoV-19/England/MILK-AB801C/2020\|EPI_ISL_623461\|20 | ···················· | ···················· | ······················ | ···················· | ···················· | ···················· |
| P | 442 | hCoV-19/England/MILK-AB7AC4/2020\|EPI_ISL_623462\|20 | ···················· | ···················· | ······················ | ···················· | ···················· | ···················· |
| P | 443 | hCoV-19/England/MILK-AB8340/2020\|EPI_ISL_623464\|20 | ···················· | ···················· | ······················ | ···················· | ···················· | ···················· |
| P | 444 | hCoV-19/England/MILK-ABB43B/2020\|EPI_ISL_623465\|20 | ···················· | ···················· | ······················ | ···················· | ···················· | ···················· |
| P | 445 | hCoV-19/England/MILK-AB8665/2020\|EPI_ISL_623466\|20 | ···················· | ···················· | ······················ | ···················· | ···················· | ···················· |
| P | 446 | hCoV-19/England/MILK-AB8726/2020\|EPI_ISL_623467\|20 | ···················· | ···················· | ······················ | ···················· | ···················· | ···················· |
| P | 447 | hCoV-19/England/MILK-AB7806/2020\|EPI_ISL_623469\|20 | ···················· | ···················· | ······················ | ···················· | ···················· | ···················· |
| P | 448 | hCoV-19/England/ALDP-A95CA8/2020\|EPI_ISL_624700\|20 | ···················· | ···················· | ······················ | ···················· | ···················· | ···················· |
| P | 449 | hCoV-19/England/ALDP-A94140/2020\|EPI_ISL_624701\|20 | ···················· | ···················· | ······················ | ···················· | ···················· | ···················· |
| P | 450 | hCoV-19/England/ALDP-A94A54/2020\|EPI_ISL_624702\|20 | ···················· | ···················· | ······················ | ···················· | ···················· | ···················· |
| P | 451 | hCoV-19/England/ALDP-A960F6/2020\|EPI_ISL_624703\|20 | ···················· | ···················· | ······················ | ···················· | ···················· | ···················· |
| P | 452 | hCoV-19/England/ALDP-A953FE/2020\|EPI_ISL_624704\|20 | ···················· | ···················· | ······················ | ···················· | ···················· | ···················· |
| P | 453 | hCoV-19/England/ALDP-A944B0/2020\|EPI_ISL_624705\|20 | ···················· | ···················· | ······················ | ···················· | ···················· | ···················· |
| P | 454 | hCoV-19/England/ALDP-A94B42/2020\|EPI_ISL_624706\|20 | ···················· | ···················· | ······················ | ···················· | ···················· | ···················· |
| P | 455 | hCoV-19/England/ALDP-A95376/2020\|EPI_ISL_624707\|20 | ···················· | ···················· | ······················ | ···················· | ···················· | ···················· |
| P | 456 | hCoV-19/England/ALDP-A94C9A/2020\|EPI_ISL_624708\|20 | ···················· | ·A·················· | ······················ | ···················· | ···················· | ···A················ |
| P | 457 | hCoV-19/England/ALDP-A95ACC/2020\|EPI_ISL_624709\|20 | ···················· | ···················· | ······················ | ···················· | ···················· | ···················· |
| P | 458 | hCoV-19/England/ALDP-A9616C/2020\|EPI_ISL_624710\|20 | ···················· | ···················· | ······················ | ···················· | ···················· | ···················· |
| P | 459 | hCoV-19/England/ALDP-A961A8/2020\|EPI_ISL_624711\|20 | ···················· | ···················· | ······················ | ···················· | ···················· | ···················· |
| P | 460 | hCoV-19/England/ALDP-A9632A/2020\|EPI_ISL_624712\|20 | ···················· | ···················· | ······················ | ···················· | ······T············· | ···················· |
| P | 461 | hCoV-19/England/ALDP-A94CC7/2020\|EPI_ISL_624713\|20 | ···················· | ···················· | ······················ | ···················· | ···················· | ···················· |
| P | 462 | hCoV-19/England/ALDP-A9609C/2020\|EPI_ISL_624716\|20 | ···················· | ···················· | ······················ | ···················· | ···················· | ···················· |
| P | 463 | hCoV-19/England/ALDP-A95A53/2020\|EPI_ISL_624717\|20 | ···················· | ···················· | ······················ | ···················· | ···················· | ···················· |
| P | 464 | hCoV-19/England/ALDP-A94FBF/2020\|EPI_ISL_624718\|20 | ···················· | ···················· | ······················ | ···················· | ···················· | ···················· |
| P | 465 | hCoV-19/England/ALDP-A9614E/2020\|EPI_ISL_624719\|20 | ···················· | ···················· | ······················ | ···················· | ···················· | ···················· |
| P | 466 | hCoV-19/England/ALDP-A95DF0/2020\|EPI_ISL_624720\|20 | ···················· | ···················· | ······················ | ···················· | ···················· | ···················· |
| P | 467 | hCoV-19/England/ALDP-A95A9F/2020\|EPI_ISL_624721\|20 | ···················· | ···················· | ······················ | ···················· | ···················· | ···················· |
| P | 468 | hCoV-19/England/ALDP-A95EB1/2020\|EPI_ISL_624722\|20 | ···················· | ···················· | ······················ | ···················· | ···················· | ···················· |
| P | 469 | hCoV-19/England/ALDP-A9620F/2020\|EPI_ISL_624723\|20 | ···················· | ···················· | ······················ | ···················· | ···················· | ···················· |
| P | 470 | hCoV-19/England/ALDP-A95E84/2020\|EPI_ISL_624725\|20 | ···················· | ···················· | ······················ | ···················· | ···················· | ···················· |
| P | 471 | hCoV-19/England/ALDP-A94605/2020\|EPI_ISL_624726\|20 | ···················· | ·········T·········· | ······················ | ···················· | ···················· | ···················· |
| P | 472 | hCoV-19/England/ALDP-A95419/2020\|EPI_ISL_624727\|20 | ···················· | ···················· | ······················ | ···················· | ···················· | ···················· |
| P | 473 | hCoV-19/England/ALDP-A95E1B/2020\|EPI_ISL_624729\|20 | ···················· | ···················· | ······················ | ···················· | ···················· | ···················· |
| P | 474 | hCoV-19/England/ALDP-A96472/2020\|EPI_ISL_624730\|20 | ···················· | ···················· | ······················ | ···················· | ···················· | ···················· |
| P | 475 | hCoV-19/England/ALDP-A94C7C/2020\|EPI_ISL_624731\|20 | ···················· | ···················· | ······················ | ···················· | ···················· | ···················· |
| P | 476 | hCoV-19/England/ALDP-A94E0D/2020\|EPI_ISL_624732\|20 | ···················· | ···················· | ······················ | ···················· | ···················· | ···················· |
| P | 477 | hCoV-19/England/ALDP-A96339/2020\|EPI_ISL_624733\|20 | ···················· | ···················· | ······················ | ···················· | ···················· | ···················· |
| P | 478 | hCoV-19/England/ALDP-A94B51/2020\|EPI_ISL_624734\|20 | ···················· | ···················· | ······················ | ···················· | ···················· | ···················· |
| P | 479 | hCoV-19/England/ALDP-A94A36/2020\|EPI_ISL_624735\|20 | ···················· | ···················· | ······················ | ···················· | ···················· | ···················· |
| P | 480 | hCoV-19/England/ALDP-A96269/2020\|EPI_ISL_624759\|20 | ···················· | ···················· | ······················ | ···················· | ···················· | ···················· |
| P | 481 | hCoV-19/England/ALDP-A964FA/2020\|EPI_ISL_624760\|20 | ···················· | ···················· | ······················ | ···················· | ······T············· | ···················· |
| P | 482 | hCoV-19/England/ALDP-A95367/2020\|EPI_ISL_624762\|20 | ···················· | ···················· | ······················ | ···················· | ···················· | ···················· |
| P | 483 | hCoV-19/England/ALDP-A944DE/2020\|EPI_ISL_624763\|20 | ···················· | ···················· | ······················ | ···················· | ···················· | ···················· |
| P | 484 | hCoV-19/England/ALDP-A9624B/2020\|EPI_ISL_624764\|20 | ···················· | ···················· | ······················ | ···················· | ···················· | ···················· |
| P | 485 | hCoV-19/England/ALDP-A95A71/2020\|EPI_ISL_624765\|20 | ···················· | ···················· | ······················ | ···················· | ···················· | ···················· |
| P | 486 | hCoV-19/England/ALDP-A9580E/2020\|EPI_ISL_624766\|20 | ···················· | ···················· | ······················ | ···················· | ···················· | ···················· |
| P | 487 | hCoV-19/England/ALDP-A95622/2020\|EPI_ISL_624767\|20 | ···················· | ···················· | ······················ | ···················· | ···················· | ···················· |
| P | 488 | hCoV-19/England/ALDP-A9424D/2020\|EPI_ISL_624768\|20 | ···················· | ···················· | ······················ | ···················· | ···················· | ···················· |
| P | 489 | hCoV-19/England/ALDP-A9622D/2020\|EPI_ISL_624769\|20 | ···················· | ···················· | ······················ | ···················· | ···················· | ···················· |
| P | 490 | hCoV-19/England/ALDP-A962E1/2020\|EPI_ISL_624771\|20 | ···················· | ···················· | ······················ | ···················· | ···················· | ···················· |
| P | 491 | hCoV-19/England/ALDP-A94A63/2020\|EPI_ISL_624772\|20 | ···················· | ···················· | ······················ | ···················· | ···················· | ···················· |
| P | 492 | hCoV-19/England/ALDP-A94F46/2020\|EPI_ISL_624773\|20 | ···················· | ···················· | ······················ | ···················· | ···················· | ···················· |
| P | 493 | hCoV-19/England/ALDP-A961B7/2020\|EPI_ISL_624774\|20 | ···················· | ···················· | ······················ | ···················· | ···················· | ···················· |
| P | 494 | hCoV-19/Wales/ALDP-A956B9/2020\|EPI_ISL_624776\|2020 | ···················· | ···················· | ······················ | ···················· | ···················· | ···················· |
| P | 495 | hCoV-19/England/ALDP-A9532B/2020\|EPI_ISL_624777\|20 | ···················· | ···················· | ······················ | ···················· | ···················· | ···················· |
| P | 496 | hCoV-19/England/ALDP-A95F18/2020\|EPI_ISL_624779\|20 | ···················· | ···················· | ······················ | ···················· | ···················· | ···················· |
| P | 497 | hCoV-19/England/ALDP-A95428/2020\|EPI_ISL_624780\|20 | ···················· | ·A·················· | ······················ | ···················· | ···················· | ···················· |
| P | 498 | hCoV-19/England/ALDP-A94C5E/2020\|EPI_ISL_624781\|20 | ···················· | ···················· | ······················ | ···················· | ···················· | ···················· |
| P | 499 | hCoV-19/England/ALDP-A94465/2020\|EPI_ISL_624782\|20 | ···················· | ···················· | ······················ | ···················· | ···················· | ···················· |
| P | 500 | hCoV-19/England/ALDP-A95A35/2020\|EPI_ISL_624783\|20 | ···················· | ···················· | ······················ | ···················· | ···················· | ···················· |
| P | 501 | hCoV-19/England/ALDP-A94508/2020\|EPI_ISL_624784\|20 | ···················· | ···················· | ······················ | ···················· | ···················· | ···················· |
| P | 502 | hCoV-19/England/ALDP-A9584A/2020\|EPI_ISL_624785\|20 | ···················· | ···················· | ······················ | ···················· | ···················· | ···················· |
| P | 503 | hCoV-19/England/ALDP-A9613F/2020\|EPI_ISL_624786\|20 | ···················· | ···················· | ······················ | ···················· | ···················· | ···················· |
| P | 504 | hCoV-19/England/ALDP-A95D2D/2020\|EPI_ISL_624787\|20 | ···················· | ···················· | ······················ | ···················· | ···················· | ···················· |
| P | 505 | hCoV-19/England/ALDP-A960AB/2020\|EPI_ISL_624789\|20 | ···················· | ···················· | ······················ | ···················· | ···················· | ···················· |
| P | 506 | hCoV-19/England/ALDP-A94F37/2020\|EPI_ISL_624790\|20 | ···················· | ···················· | ······················ | ···················· | ···················· | ···················· |
| P | 507 | hCoV-19/England/ALDP-A9478A/2020\|EPI_ISL_624791\|20 | ···················· | ·A·················· | ······················ | ···················· | ···················· | ···A················ |
| P | 508 | hCoV-19/England/ALDP-A95EEE/2020\|EPI_ISL_624792\|20 | ···················· | ···················· | ······················ | ···················· | ···················· | ···················· |
| P | 509 | hCoV-19/England/ALDP-A95895/2020\|EPI_ISL_624793\|20 | ···················· | ···················· | ······················ | ···················· | ···················· | ···················· |
| P | 510 | hCoV-19/England/ALDP-A94B24/2020\|EPI_ISL_624795\|20 | ···················· | ···················· | ······················ | ···················· | ···················· | ···················· |
| P | 511 | hCoV-19/England/ALDP-A95349/2020\|EPI_ISL_624796\|20 | ···················· | ···················· | ······················ | ···················· | ···················· | ···················· |
| P | 512 | hCoV-19/England/ALDP-A96014/2020\|EPI_ISL_624797\|20 | ···················· | ···················· | ······················ | ···················· | ···················· | ···················· |
| P | 513 | hCoV-19/Denmark/DCGC-3456/2020\|EPI_ISL_616220\|2020 | ···················· | ···················· | ······················ | ···················· | ···················· | ···················· |

| | | | | | | | |
|---|---|---|---|---|---|---|---|
| P | 514 hCoV-19/Denmark/DCGC-3457/2020\|EPI_ISL_616221\|2020 | ···················· | ···················· | ····················· | ···················· | ···················· | ···················· |
| P | 515 hCoV-19/Denmark/DCGC-3458/2020\|EPI_ISL_616222\|2020 | ···················· | ·A·················· | ····················· | ···················· | ···················· | ··A················· |
| P | 516 hCoV-19/Denmark/DCGC-3459/2020\|EPI_ISL_616223\|2020 | ···················· | ···················· | ····················· | ···················· | ···················· | ···················· |
| P | 517 hCoV-19/Denmark/DCGC-3460/2020\|EPI_ISL_616224\|2020 | ···················· | ···················· | ····················· | ···················· | ···················· | ···················· |
| P | 518 hCoV-19/Denmark/DCGC-3584/2020\|EPI_ISL_616225\|2020 | ···················· | ···················· | ····················· | ···················· | ···················· | ···················· |
| P | 519 hCoV-19/Denmark/DCGC-6296/2020\|EPI_ISL_616226\|2020 | ···················· | ···················· | ····················· | ···················· | ···················· | ···················· |
| P | 520 hCoV-19/Denmark/DCGC-3585/2020\|EPI_ISL_616227\|2020 | ···················· | ···················· | ····················· | ···················· | ···················· | ···················· |
| P | 521 hCoV-19/Denmark/DCGC-6297/2020\|EPI_ISL_616228\|2020 | ···················· | ···················· | ····················· | ···················· | ···················· | ···················· |
| P | 522 hCoV-19/Denmark/DCGC-3587/2020\|EPI_ISL_616230\|2020 | ···················· | ···················· | ····················· | ···················· | ···················· | ···················· |
| P | 523 hCoV-19/Denmark/DCGC-3588/2020\|EPI_ISL_616231\|2020 | ···················· | ···················· | ····················· | ···················· | ···················· | ···················· |
| P | 524 hCoV-19/Denmark/DCGC-3593/2020\|EPI_ISL_616236\|2020 | ···················· | ·A·················· | ····················· | ···················· | ···················· | ···················· |
| P | 525 hCoV-19/Denmark/DCGC-3594/2020\|EPI_ISL_616237\|2020 | ···················· | ···················· | ····················· | ···················· | ···················· | ···················· |
| P | 526 hCoV-19/Denmark/DCGC-3596/2020\|EPI_ISL_616239\|2020 | ···················· | ···················· | ····················· | ···················· | ···················· | ···················· |
| P | 527 hCoV-19/Denmark/DCGC-2199/2020\|EPI_ISL_617500\|2020 | ···················· | ···················· | ····················· | ···················· | ···················· | ···················· |
| P | 528 hCoV-19/Denmark/DCGC-2200/2020\|EPI_ISL_617501\|2020 | ···················· | ···················· | ····················· | ···················· | ···················· | ···················· |
| P | 529 hCoV-19/Denmark/DCGC-2201/2020\|EPI_ISL_617502\|2020 | ···················· | ···················· | ····················· | ···················· | ···················· | ···················· |
| P | 530 hCoV-19/Denmark/DCGC-2202/2020\|EPI_ISL_617503\|2020 | ···················· | ·A·················· | ····················· | ···················· | ···················· | ···················· |
| P | 531 hCoV-19/Denmark/DCGC-2204/2020\|EPI_ISL_617505\|2020 | ···················· | ···················· | ····················· | ···················· | ···················· | ···················· |
| P | 532 hCoV-19/Denmark/DCGC-2205/2020\|EPI_ISL_617506\|2020 | ···················· | ···················· | ····················· | ···················· | ···················· | ···················· |
| P | 533 hCoV-19/Denmark/DCGC-2425/2020\|EPI_ISL_617507\|2020 | ···················· | ·A·················· | ····················· | ···················· | ···················· | ···················· |
| P | 534 hCoV-19/Denmark/DCGC-2245/2020\|EPI_ISL_617550\|2020 | ···················· | ···················· | ····················· | ···················· | ···················· | ···················· |
| P | 535 hCoV-19/Denmark/DCGC-2246/2020\|EPI_ISL_617551\|2020 | ···················· | ···················· | ····················· | ···················· | ···················· | ···················· |
| P | 536 hCoV-19/Denmark/DCGC-2247/2020\|EPI_ISL_617552\|2020 | ···················· | ···················· | ····················· | ···················· | ···················· | ···················· |
| P | 537 hCoV-19/Denmark/DCGC-2248/2020\|EPI_ISL_617553\|2020 | ···················· | ···················· | ····················· | ···················· | ···················· | ···················· |
| P | 538 hCoV-19/Denmark/DCGC-2249/2020\|EPI_ISL_617554\|2020 | ···················· | ···················· | ····················· | ···················· | ···················· | ···················· |
| P | 539 hCoV-19/Denmark/DCGC-2375/2020\|EPI_ISL_617555\|2020 | ···················· | ···················· | ····················· | ···················· | ···················· | ···················· |
| P | 540 hCoV-19/Denmark/DCGC-2376/2020\|EPI_ISL_617556\|2020 | ···················· | ···················· | ····················· | ···················· | ···················· | ···················· |
| P | 541 hCoV-19/Denmark/DCGC-2377/2020\|EPI_ISL_617557\|2020 | ···················· | ···················· | ····················· | ···················· | ···················· | ···················· |
| P | 542 hCoV-19/Denmark/DCGC-2378/2020\|EPI_ISL_617558\|2020 | ···················· | ···················· | ···········T········· | ···················· | ···················· | ···················· |
| P | 543 hCoV-19/Denmark/DCGC-2379/2020\|EPI_ISL_617559\|2020 | ···················· | ···················· | ····················· | ···················· | ···················· | ···················· |
| P | 544 hCoV-19/Denmark/DCGC-2383/2020\|EPI_ISL_617563\|2020 | ···················· | ···················· | ····················· | ···················· | ···················· | ···················· |
| P | 545 hCoV-19/Denmark/DCGC-2385/2020\|EPI_ISL_617565\|2020 | ···················· | ···················· | ····················· | ···················· | ···················· | ···················· |
| P | 546 hCoV-19/Denmark/DCGC-2386/2020\|EPI_ISL_617566\|2020 | ···················· | ···················· | ····················· | ···················· | ···················· | ···················· |
| P | 547 hCoV-19/Denmark/DCGC-2387/2020\|EPI_ISL_617567\|2020 | ···················· | ···················· | ····················· | ···················· | ···················· | ···················· |
| P | 548 hCoV-19/Denmark/DCGC-2388/2020\|EPI_ISL_617568\|2020 | ···················· | ···················· | ····················· | ···················· | ···················· | ···················· |
| P | 549 hCoV-19/Denmark/DCGC-2389/2020\|EPI_ISL_617569\|2020 | ···················· | ···················· | ····················· | ···················· | ···················· | ···················· |
| P | 550 hCoV-19/Denmark/DCGC-1344/2020\|EPI_ISL_618801\|2020 | ···················· | ···················· | ····················· | ···················· | ···················· | ···················· |
| P | 551 hCoV-19/Denmark/DCGC-1352/2020\|EPI_ISL_618802\|2020 | ···················· | ···················· | ····················· | ···················· | ···················· | ···················· |
| P | 552 hCoV-19/Denmark/DCGC-1353/2020\|EPI_ISL_618803\|2020 | ···················· | ···················· | ····················· | ···················· | ···················· | ···················· |
| P | 553 hCoV-19/Denmark/DCGC-1354/2020\|EPI_ISL_618804\|2020 | ···················· | ···················· | ····················· | ···················· | ···················· | ···················· |
| P | 554 hCoV-19/Denmark/DCGC-1386/2020\|EPI_ISL_618805\|2020 | ···················· | ···················· | ····················· | ···················· | ···················· | ···················· |
| P | 555 hCoV-19/Denmark/DCGC-1387/2020\|EPI_ISL_618806\|2020 | ···················· | ···················· | ····················· | ···················· | ···················· | ···················· |
| P | 556 hCoV-19/Denmark/DCGC-1732/2020\|EPI_ISL_618807\|2020 | ···················· | ···················· | ····················· | ···················· | ···················· | ···················· |
| P | 557 hCoV-19/Denmark/DCGC-1679/2020\|EPI_ISL_618808\|2020 | ···················· | ···················· | ····················· | ···················· | ···················· | ···················· |
| P | 558 hCoV-19/Denmark/DCGC-7856/2020\|EPI_ISL_618809\|2020 | ···················· | ···················· | ····················· | ···················· | ···················· | ···················· |
| P | 559 hCoV-19/Denmark/DCGC-7871/2020\|EPI_ISL_618811\|2020 | ···················· | ·A·················· | ····················· | ···················· | ···················· | ···················· |
| P | 560 hCoV-19/Denmark/DCGC-7882/2020\|EPI_ISL_618812\|2020 | ···················· | ···················· | ····················· | ···················· | ···················· | ···················· |
| P | 561 hCoV-19/Denmark/DCGC-7890/2020\|EPI_ISL_618813\|2020 | ···················· | ···················· | ····················· | ···················· | ···················· | ···················· |
| P | 562 hCoV-19/Denmark/DCGC-7899/2020\|EPI_ISL_618814\|2020 | ···················· | ···················· | ····················· | ···················· | ···················· | ···················· |
| P | 563 hCoV-19/Denmark/DCGC-7905/2020\|EPI_ISL_618815\|2020 | ···················· | ···················· | ····················· | ···················· | ···················· | ···················· |
| P | 564 hCoV-19/Denmark/DCGC-7918/2020\|EPI_ISL_618817\|2020 | ···················· | ···················· | ····················· | ···················· | ···················· | ···················· |
| P | 565 hCoV-19/Denmark/DCGC-7854/2020\|EPI_ISL_618818\|2020 | ···················· | ···················· | ····················· | ···················· | ···················· | ···················· |
| P | 566 hCoV-19/Denmark/DCGC-7857/2020\|EPI_ISL_618819\|2020 | ···················· | ···················· | ····················· | ···················· | ···················· | ···················· |
| P | 567 hCoV-19/Denmark/DCGC-7864/2020\|EPI_ISL_618820\|2020 | ···················· | ···················· | ····················· | ···················· | ···················· | ···················· |
| P | 568 hCoV-19/Denmark/DCGC-7872/2020\|EPI_ISL_618821\|2020 | ···················· | ·N·················· | ····················· | ···················· | ···················· | ··A················· |
| P | 569 hCoV-19/Denmark/DCGC-7877/2020\|EPI_ISL_618822\|2020 | ···················· | ·A·················· | ····················· | ···················· | ···················· | ···················· |
| P | 570 hCoV-19/Denmark/DCGC-7883/2020\|EPI_ISL_618823\|2020 | ···················· | ···················· | ····················· | ···················· | ···················· | ···················· |
| P | 571 hCoV-19/Denmark/DCGC-7891/2020\|EPI_ISL_618824\|2020 | ···················· | ···················· | ····················· | ···················· | ···················· | ···················· |
| P | 572 hCoV-19/Denmark/DCGC-7900/2020\|EPI_ISL_618825\|2020 | ···················· | ···················· | ····················· | ···················· | ···················· | ···················· |
| P | 573 hCoV-19/Denmark/DCGC-7919/2020\|EPI_ISL_618827\|2020 | ···················· | ···················· | ····················· | ···················· | ···················· | ···················· |
| P | 574 hCoV-19/Denmark/DCGC-7865/2020\|EPI_ISL_618829\|2020 | ···················· | ···················· | ····················· | ···················· | ···················· | ···················· |
| P | 575 hCoV-19/Denmark/DCGC-7878/2020\|EPI_ISL_618830\|2020 | ···················· | ···················· | ····················· | ···················· | ···················· | ···················· |
| P | 576 hCoV-19/Denmark/DCGC-7884/2020\|EPI_ISL_618831\|2020 | ···················· | ···················· | ····················· | ···················· | ···················· | ···················· |
| P | 577 hCoV-19/Denmark/DCGC-7892/2020\|EPI_ISL_618832\|2020 | ···················· | ···················· | ····················· | ···················· | ···················· | ···················· |
| P | 578 hCoV-19/Denmark/DCGC-7896/2020\|EPI_ISL_618833\|2020 | ···················· | ···················· | ····················· | ···················· | ···················· | ···················· |
| P | 579 hCoV-19/Denmark/DCGC-7906/2020\|EPI_ISL_618834\|2020 | ···················· | ···················· | ····················· | ···················· | ···················· | ···················· |
| P | 580 hCoV-19/Denmark/DCGC-7859/2020\|EPI_ISL_618835\|2020 | ···················· | ···················· | ····················· | ···················· | ···················· | ···················· |
| P | 581 hCoV-19/Denmark/DCGC-7866/2020\|EPI_ISL_618836\|2020 | ···················· | ···················· | ····················· | ···················· | ···················· | ···················· |
| P | 582 hCoV-19/Denmark/DCGC-7873/2020\|EPI_ISL_618837\|2020 | ···················· | ···················· | ····················· | ···················· | ···················· | ···················· |
| P | 583 hCoV-19/Denmark/DCGC-7885/2020\|EPI_ISL_618838\|2020 | ···················· | ···················· | ····················· | ···················· | ···················· | ···················· |
| P | 584 hCoV-19/Denmark/DCGC-7893/2020\|EPI_ISL_618839\|2020 | ···················· | ···················· | ····················· | ···················· | ···················· | ···················· |
| P | 585 hCoV-19/Denmark/DCGC-7921/2020\|EPI_ISL_618880\|2020 | ···················· | ···················· | ····················· | ···················· | ···················· | ···················· |
| P | 586 hCoV-19/Denmark/DCGC-7927/2020\|EPI_ISL_618881\|2020 | ···················· | ···················· | ····················· | ···················· | ···················· | ···················· |
| P | 587 hCoV-19/Denmark/DCGC-7933/2020\|EPI_ISL_618882\|2020 | ···················· | ···················· | ····················· | ···················· | ···················· | ···················· |
| P | 588 hCoV-19/Denmark/DCGC-7945/2020\|EPI_ISL_618884\|2020 | ···················· | ···················· | ····················· | ···················· | ···················· | ···················· |
| P | 589 hCoV-19/Denmark/DCGC-7952/2020\|EPI_ISL_618885\|2020 | ···················· | ···················· | ····················· | ···················· | ···················· | ···················· |
| P | 590 hCoV-19/Denmark/DCGC-7958/2020\|EPI_ISL_618886\|2020 | ···················· | ···················· | ····················· | ···················· | ···················· | ···················· |
| P | 591 hCoV-19/Denmark/DCGC-7966/2020\|EPI_ISL_618887\|2020 | ···················· | ···················· | ····················· | ···················· | ···················· | ···················· |
| P | 592 hCoV-19/Denmark/DCGC-7928/2020\|EPI_ISL_618888\|2020 | ···················· | ···················· | ····················· | ···················· | ···················· | ···················· |
| P | 593 hCoV-19/Denmark/DCGC-7967/2020\|EPI_ISL_618889\|2020 | ···················· | ···················· | ····················· | ···················· | ···················· | ···················· |

P 594 hCoV-19/Denmark/DCGC-7948/2020|EPI_ISL_618894|2020 ···················· ···················· ···················· ···················· ···················· ····················
P 595 hCoV-19/Denmark/DCGC-7959/2020|EPI_ISL_618895|2020 ···················· ···················· ···················· ···················· ···················· ····················
P 596 hCoV-19/Denmark/DCGC-7968/2020|EPI_ISL_618896|2020 ···················· ···················· ···················· ···················· ···················· ····················
P 597 hCoV-19/Denmark/DCGC-7922/2020|EPI_ISL_618897|2020 ···················· ···················· ···················· ···················· ···················· ····················
P 598 hCoV-19/Denmark/DCGC-7929/2020|EPI_ISL_618898|2020 ···················· ···················· ···················· ···················· ···················· ····················
P 599 hCoV-19/Denmark/DCGC-7938/2020|EPI_ISL_618899|2020 ···················· ···················· ···················· ···················· ···················· ····················
P 600 hCoV-19/Denmark/DCGC-4063/2020|EPI_ISL_622019|2020 ···················· ···················· ···················· ···················· ···················· ·········C··········
P 601 hCoV-19/Denmark/DCGC-4064/2020|EPI_ISL_622020|2020 ···················· ···················· ···················· ···················· ···················· ····················
P 602 hCoV-19/Denmark/DCGC-4065/2020|EPI_ISL_622021|2020 ···················· ···················· ···················· ···················· ···················· ····················
P 603 hCoV-19/Denmark/DCGC-4066/2020|EPI_ISL_622022|2020 ···················· ···················· ···················· ···················· ···················· ····················
P 604 hCoV-19/Denmark/DCGC-4068/2020|EPI_ISL_622024|2020 ···················· ···················· ···················· ···················· ···················· ····················
P 605 hCoV-19/Denmark/DCGC-4069/2020|EPI_ISL_622025|2020 ···················· ···················· ···················· ···················· ···················· ····················
P 606 hCoV-19/Denmark/DCGC-4070/2020|EPI_ISL_622026|2020 ···················· ···················· ···················· ···················· ···················· ····················
P 607 hCoV-19/Denmark/DCGC-4071/2020|EPI_ISL_622027|2020 ···················· ···················· ···················· ···················· ···················· ····················
P 608 hCoV-19/Denmark/DCGC-4072/2020|EPI_ISL_622028|2020 ···················· ···················· ···················· ···················· ···················· ····················
P 609 hCoV-19/Denmark/DCGC-4074/2020|EPI_ISL_622030|2020 ···················· ···················· ···················· ···················· ···················· ····················
P 610 hCoV-19/Denmark/DCGC-4075/2020|EPI_ISL_622031|2020 ···················· ···················· ···················· ···················· ···················· ····················
P 611 hCoV-19/Denmark/DCGC-4076/2020|EPI_ISL_622032|2020 ···················· ···················· ···················· ···················· ···················· ····················
P 612 hCoV-19/Denmark/DCGC-4077/2020|EPI_ISL_622033|2020 ···················· ···················· ···················· ···················· ···················· ····················
P 613 hCoV-19/Denmark/DCGC-4078/2020|EPI_ISL_622034|2020 ···················· ···················· ···················· ···················· ···················· ····················
P 614 hCoV-19/Denmark/DCGC-4079/2020|EPI_ISL_622035|2020 ···················· ···················· ···················· ···················· ···················· ····················
P 615 hCoV-19/Denmark/DCGC-4080/2020|EPI_ISL_622036|2020 ···················· ···················· ···················· ···················· ···················· ····················
P 616 hCoV-19/Denmark/DCGC-4081/2020|EPI_ISL_622037|2020 ···················· ···················· ···················· ···················· ···················· ····················
P 617 hCoV-19/Denmark/DCGC-4082/2020|EPI_ISL_622038|2020 ···················· ···················· ···················· ···················· ···················· ····················
P 618 hCoV-19/Denmark/DCGC-4083/2020|EPI_ISL_622039|2020 ···················· ···················· ···················· ···················· ···················· ····················
P 619 hCoV-19/Denmark/DCGC-4084/2020|EPI_ISL_622040|2020 ···················· ···················· ···················· ···················· ···················· ····················
P 620 hCoV-19/Denmark/DCGC-4085/2020|EPI_ISL_622041|2020 ···················· ···················· ···················· ···················· ···················· ····················
P 621 hCoV-19/Denmark/DCGC-4086/2020|EPI_ISL_622042|2020 ···················· ···················· ···················· ···················· ···················· ····················
P 622 hCoV-19/Denmark/DCGC-4087/2020|EPI_ISL_622043|2020 ···················· ···················· ···················· ···················· ···················· ····················
P 623 hCoV-19/Denmark/DCGC-4088/2020|EPI_ISL_622044|2020 ···················· ···················· ···················· ···················· ···················· ····················
P 624 hCoV-19/Denmark/DCGC-4089/2020|EPI_ISL_622045|2020 ···················· ···················· ···················· ···················· ···················· ····················
P 625 hCoV-19/Denmark/DCGC-4090/2020|EPI_ISL_622046|2020 ···················· ···················· ···················· ···················· ···················· ····················
P 626 hCoV-19/Denmark/DCGC-4091/2020|EPI_ISL_622047|2020 ···················· ···················· ···················· ···················· ···················· ····················
P 627 hCoV-19/Denmark/DCGC-4092/2020|EPI_ISL_622048|2020 ···················· ···················· ···················· ···················· ···················· ····················
P 628 hCoV-19/Denmark/DCGC-4093/2020|EPI_ISL_622049|2020 ···················· ···················· ···················· ···················· ···················· ····················
P 629 hCoV-19/Denmark/DCGC-4094/2020|EPI_ISL_622050|2020 ···················· ···················· ···················· ···················· ···················· ····················
P 630 hCoV-19/Denmark/DCGC-4095/2020|EPI_ISL_622051|2020 ···················· ···················· ···················· ···················· ···················· ····················
P 631 hCoV-19/Denmark/DCGC-4096/2020|EPI_ISL_622052|2020 ···················· ···················· ···················· ···················· ···················· ····················
P 632 hCoV-19/Denmark/DCGC-4097/2020|EPI_ISL_622053|2020 ···················· ···················· ···················· ···················· ···················· ····················
P 633 hCoV-19/Denmark/DCGC-4098/2020|EPI_ISL_622054|2020 ···················· ···················· ···················· ···················· ···················· ····················
P 634 hCoV-19/Denmark/DCGC-4099/2020|EPI_ISL_622055|2020 ···················· ···················· ···················· ···················· ···················· ····················
P 635 hCoV-19/Denmark/DCGC-4100/2020|EPI_ISL_622056|2020 ···················· ···················· ···················· ···················· ···················· ····················
P 636 hCoV-19/Denmark/DCGC-4103/2020|EPI_ISL_622059|2020 ···················· ···················· ···················· ···················· ···················· ····················
P 637 hCoV-19/Denmark/DCGC-4104/2020|EPI_ISL_622060|2020 ···················· ···················· ···················· ···················· ···················· ····················
P 638 hCoV-19/Denmark/DCGC-4106/2020|EPI_ISL_622062|2020 ···················· ···················· ···················· ···················· ···················· ····················
P 639 hCoV-19/Denmark/DCGC-4107/2020|EPI_ISL_622063|2020 ···················· ···················· ···················· ···················· ···················· ····················
P 640 hCoV-19/Denmark/DCGC-4108/2020|EPI_ISL_622064|2020 ···················· ···················· ···················· ···················· ···················· ····················
P 641 hCoV-19/Denmark/DCGC-4110/2020|EPI_ISL_622066|2020 ···················· ···················· ···················· ···················· ···················· ····················
P 642 hCoV-19/Denmark/DCGC-4111/2020|EPI_ISL_622067|2020 ···················· ···················· ···················· ···················· ···················· ····················
P 643 hCoV-19/Denmark/DCGC-4112/2020|EPI_ISL_622068|2020 ···················· ···················· ···················· ···················· ···················· ····················
P 644 hCoV-19/Denmark/DCGC-4114/2020|EPI_ISL_622070|2020 ···················· ···················· ···················· ···················· ···················· ····················
P 645 hCoV-19/Denmark/DCGC-4115/2020|EPI_ISL_622071|2020 ···················· ···················· ···················· ···················· ···················· ····················
P 646 hCoV-19/Denmark/DCGC-4116/2020|EPI_ISL_622072|2020 ···················· ···················· ···················· ···················· ···················· ····················
P 647 hCoV-19/Denmark/DCGC-4117/2020|EPI_ISL_622073|2020 ···················· ···················· ···················· ···················· ···················· ····················
P 648 hCoV-19/Denmark/DCGC-4118/2020|EPI_ISL_622074|2020 ···················· ···················· ···················· ···················· ···················· ····················
P 649 hCoV-19/Denmark/DCGC-4119/2020|EPI_ISL_622075|2020 ···················· ···················· ···················· ···················· ···················· ····················
P 650 hCoV-19/Denmark/DCGC-4121/2020|EPI_ISL_622077|2020 ···················· ···················· ···················· ···················· ···················· ····················
P 651 hCoV-19/Denmark/DCGC-4185/2020|EPI_ISL_622078|2020 ···················· ···················· ···················· ···················· ···················· ····················
P 652 hCoV-19/Denmark/DCGC-4186/2020|EPI_ISL_622079|2020 ···················· ···················· ···················· ···················· ···················· ····················
P 653 hCoV-19/Denmark/DCGC-4187/2020|EPI_ISL_622080|2020 ···················· ···················· ···················· ···················· ···················· ····················
P 654 hCoV-19/Denmark/DCGC-4188/2020|EPI_ISL_622081|2020 ···················· ···················· ···················· ···················· ···················· ····················
P 655 hCoV-19/Denmark/DCGC-4189/2020|EPI_ISL_622082|2020 ···················· ···················· ···················· ···················· ···················· ····················
P 656 hCoV-19/Denmark/DCGC-4190/2020|EPI_ISL_622083|2020 ···················· ···················· ···················· ···················· ···················· ····················
P 657 hCoV-19/Denmark/DCGC-4191/2020|EPI_ISL_622084|2020 ···················· ···················· ···················· ···················· ···················· ····················
P 658 hCoV-19/Denmark/DCGC-4192/2020|EPI_ISL_622085|2020 ···················· ···················· ···················· ···················· ···················· ····················
P 659 hCoV-19/Denmark/DCGC-4193/2020|EPI_ISL_622086|2020 ···················· ···················· ···················· ···················· ···················· ····················
P 660 hCoV-19/Denmark/DCGC-4194/2020|EPI_ISL_622087|2020 ···················· ···················· ···················· ···················· ···················· ····················
P 661 hCoV-19/Denmark/DCGC-4195/2020|EPI_ISL_622088|2020 ···················· ···················· ···················· ···················· ···················· ····················
P 662 hCoV-19/Denmark/DCGC-4196/2020|EPI_ISL_622089|2020 ···················· ···················· ···················· ···················· ···················· ····················
P 663 hCoV-19/Denmark/DCGC-4197/2020|EPI_ISL_622090|2020 ···················· ···················· ···················· ···················· ···················· ····················
P 664 hCoV-19/Denmark/DCGC-4198/2020|EPI_ISL_622091|2020 ···················· ···················· ···················· ···················· ···················· ····················
P 665 hCoV-19/Denmark/DCGC-4199/2020|EPI_ISL_622092|2020 ···················· ···················· ···················· ···················· ···················· ····················
P 666 hCoV-19/Denmark/DCGC-4200/2020|EPI_ISL_622093|2020 ···················· ···················· ···················· ···················· ···················· ····················
P 667 hCoV-19/Denmark/DCGC-4201/2020|EPI_ISL_622094|2020 ···················· ···················· ···················· ···················· ···················· ····················
P 668 hCoV-19/Denmark/DCGC-4202/2020|EPI_ISL_622095|2020 ···················· ···················· ···················· ···················· ···················· ····················
P 669 hCoV-19/Denmark/DCGC-4203/2020|EPI_ISL_622096|2020 ···················· ···················· ···················· ···················· ···················· ····················
P 670 hCoV-19/Denmark/DCGC-4204/2020|EPI_ISL_622097|2020 ···················· ···················· ···················· ···················· ···················· ····················
P 671 hCoV-19/Denmark/DCGC-4205/2020|EPI_ISL_622098|2020 ···················· ···················· ···················· ···················· ···················· ····················
P 672 hCoV-19/Denmark/DCGC-4206/2020|EPI_ISL_622099|2020 ···················· ·A·················· ···················· ···················· ···················· ··A·················
P 673 hCoV-19/England/MILK-AB84C5/2020|EPI_ISL_623350|20 ···················· ···················· ···················· ···················· ···················· ····················

| | | | | | | | | |
|---|---|---|---|---|---|---|---|---|
| P | 674 | hCoV-19/England/MILK-ABB477/2020\|EPI_ISL_623351\|20 | ···················· | ···················· | ···················· | ···················· | ···················· | ···················· |
| P | 675 | hCoV-19/England/MILK-ABB3A7/2020\|EPI_ISL_623352\|20 | ···················· | ···················· | ···················· | ···················· | ···················· | ···················· |
| P | 676 | hCoV-19/England/MILK-AB761B/2020\|EPI_ISL_623353\|20 | ···················· | ···················· | ···················· | ···················· | ···················· | ···················· |
| P | 677 | hCoV-19/England/MILK-AB7A4C/2020\|EPI_ISL_623354\|20 | ···················· | ···················· | ···················· | ···················· | ···················· | ···················· |
| P | 678 | hCoV-19/England/MILK-AB74A8/2020\|EPI_ISL_623355\|20 | ···················· | ···················· | ···················· | ···················· | ······T············· | ···················· |
| P | 679 | hCoV-19/England/MILK-AB8270/2020\|EPI_ISL_623356\|20 | ···················· | ···················· | ···················· | ···················· | ···················· | ···················· |
| P | 680 | hCoV-19/England/MILK-AB7781/2020\|EPI_ISL_623357\|20 | ···················· | ···················· | ···················· | ···················· | ···················· | ···················· |
| P | 681 | hCoV-19/England/MILK-ABB468/2020\|EPI_ISL_623358\|20 | ···················· | ···················· | ···················· | ···················· | ···················· | ···················· |
| P | 682 | hCoV-19/England/CAMC-AAFC7E/2020\|EPI_ISL_623359\|20 | ···················· | ···················· | ···················· | ···················· | ···················· | ···················· |
| P | 683 | hCoV-19/England/CAMC-AAFD4E/2020\|EPI_ISL_623360\|20 | ···················· | ···················· | ···················· | ···················· | ···················· | ···················· |
| P | 684 | hCoV-19/England/MILK-AB7DCB/2020\|EPI_ISL_623361\|20 | ···················· | ···················· | ···················· | ···················· | ······T············· | ···················· |
| P | 685 | hCoV-19/England/MILK-AB86B0/2020\|EPI_ISL_623362\|20 | ···················· | ···················· | ···················· | ···················· | ···················· | ···················· |
| P | 686 | hCoV-19/England/CAMC-AAFF0C/2020\|EPI_ISL_623363\|20 | ···················· | ···················· | ···················· | ···················· | ···················· | ···················· |
| P | 687 | hCoV-19/England/CAMC-AAFE69/2020\|EPI_ISL_623364\|20 | ···················· | ···················· | ···················· | ···················· | ···················· | ···················· |
| P | 688 | hCoV-19/England/MILK-ABB0DD/2020\|EPI_ISL_623365\|20 | ···················· | ···················· | ···················· | ···················· | ···················· | ···················· |
| P | 689 | hCoV-19/England/CAMC-AB015D/2020\|EPI_ISL_623366\|20 | ···················· | ···················· | ···················· | ···················· | ···················· | ···················· |
| P | 690 | hCoV-19/England/MILK-AB7745/2020\|EPI_ISL_623367\|20 | ···················· | ···················· | ···················· | ···················· | ···················· | ···················· |
| P | 691 | hCoV-19/England/MILK-AB803A/2020\|EPI_ISL_623368\|20 | ···················· | ···················· | ···················· | ···················· | ···················· | ···················· |
| P | 692 | hCoV-19/England/MILK-ABB5ED/2020\|EPI_ISL_623369\|20 | ···················· | ·A·················· | ···················· | ···················· | ···················· | ···················· |
| P | 693 | hCoV-19/England/CAMC-AAFB17/2020\|EPI_ISL_623370\|20 | ···················· | ···················· | ···················· | ···················· | ···················· | ···················· |
| P | 694 | hCoV-19/England/MILK-AB80B2/2020\|EPI_ISL_623372\|20 | ···················· | ···················· | ···················· | ···················· | ···················· | ···················· |
| P | 695 | hCoV-19/England/MILK-AB7499/2020\|EPI_ISL_623373\|20 | ···················· | ···················· | ···················· | ···················· | ···················· | ···················· |
| P | 696 | hCoV-19/England/MILK-ABB2D7/2020\|EPI_ISL_623375\|20 | ···················· | ···················· | ···················· | ···················· | ···················· | ···················· |
| P | 697 | hCoV-19/England/MILK-ABB7C9/2020\|EPI_ISL_623376\|20 | ···················· | ···················· | ···················· | ···················· | ···················· | ···················· |
| P | 698 | hCoV-19/England/MILK-AB76FD/2020\|EPI_ISL_623377\|20 | ···················· | ···················· | ···················· | ···················· | ···················· | ···················· |
| P | 699 | hCoV-19/England/MILK-ABB79C/2020\|EPI_ISL_623378\|20 | ···················· | ···················· | ···················· | ···················· | ···················· | ···················· |
| P | 700 | hCoV-19/England/MILK-AB84E3/2020\|EPI_ISL_623379\|20 | ···················· | ···················· | ···················· | ···················· | ···················· | ···················· |
| P | 701 | hCoV-19/England/MILK-ABB0BF/2020\|EPI_ISL_623380\|20 | ···················· | ···················· | ···················· | ···················· | ···················· | ···················· |
| P | 702 | hCoV-19/England/MILK-AB746C/2020\|EPI_ISL_623381\|20 | ···················· | ···················· | ···················· | ···················· | ···················· | ···················· |
| P | 703 | hCoV-19/England/MILK-AB748A/2020\|EPI_ISL_623382\|20 | ···················· | ···················· | ···················· | ···················· | ···················· | ···················· |
| P | 704 | hCoV-19/England/MILK-AB8322/2020\|EPI_ISL_623383\|20 | ···················· | ···················· | ···················· | ···················· | ···················· | ···················· |
| P | 705 | hCoV-19/England/MILK-AB8128/2020\|EPI_ISL_623384\|20 | ···················· | ·A·················· | ···················· | ···················· | ···················· | ··A················· |
| P | 706 | hCoV-19/England/CAMC-AB030C/2020\|EPI_ISL_623385\|20 | ···················· | ···················· | ···················· | ···················· | ···················· | ···················· |
| P | 707 | hCoV-19/England/CAMC-AAFE0F/2020\|EPI_ISL_623386\|20 | ···················· | ···················· | ···················· | ···················· | ···················· | ···················· |
| P | 708 | hCoV-19/England/MILK-AB7E40/2020\|EPI_ISL_623389\|20 | ···················· | ···················· | ···················· | ···················· | ···················· | ···················· |
| P | 709 | hCoV-19/England/CAMC-AAFB26/2020\|EPI_ISL_623390\|20 | ···················· | ···················· | ···················· | ···················· | ···················· | ···················· |
| P | 710 | hCoV-19/England/CAMC-AAFCE7/2020\|EPI_ISL_623392\|20 | ···················· | ···················· | ···················· | ···················· | ······T············· | ···················· |
| P | 711 | hCoV-19/England/CAMC-AAFD8A/2020\|EPI_ISL_623393\|20 | ···················· | ···················· | ···················· | ···················· | ···················· | ···················· |
| P | 712 | hCoV-19/England/MILK-ABB732/2020\|EPI_ISL_623394\|20 | ···················· | ·A·················· | ···················· | ···················· | ···················· | ··A················· |
| P | 713 | hCoV-19/England/MILK-AB80D0/2020\|EPI_ISL_623395\|20 | ···················· | ···················· | ···················· | ···················· | ···················· | ···················· |
| P | 714 | hCoV-19/England/MILK-AB82AD/2020\|EPI_ISL_623396\|20 | ···················· | ···················· | ···················· | ···················· | ···················· | ···················· |
| P | 715 | hCoV-19/England/CAMC-AAF968/2020\|EPI_ISL_623397\|20 | ···················· | ···················· | ···················· | ···················· | ···················· | ···················· |
| P | 716 | hCoV-19/England/CAMC-AB016C/2020\|EPI_ISL_623398\|20 | ···················· | ···················· | ···················· | ···················· | ···················· | ···················· |
| P | 717 | hCoV-19/England/MILK-AB7E31/2020\|EPI_ISL_623405\|20 | ···················· | ···················· | ···················· | ···················· | ···················· | ···················· |
| P | 718 | hCoV-19/England/MILK-AB7D8F/2020\|EPI_ISL_623406\|20 | ···················· | ···················· | ···················· | ···················· | ···················· | ···················· |
| P | 719 | hCoV-19/England/MILK-ABB3F2/2020\|EPI_ISL_623407\|20 | ···················· | ···················· | ···················· | ···················· | ······T············· | ···················· |
| P | 720 | hCoV-19/England/MILK-AB8094/2020\|EPI_ISL_623408\|20 | ···················· | ···················· | ···················· | ···················· | ······T············· | ···················· |
| P | 721 | hCoV-19/England/CAMC-AAFE2D/2020\|EPI_ISL_623409\|20 | ···················· | ···················· | ···················· | ···················· | ···················· | ···················· |
| P | 722 | hCoV-19/England/MILK-AB81FB/2020\|EPI_ISL_623410\|20 | ···················· | ···················· | ···················· | ···················· | ······T············· | ···················· |
| P | 723 | hCoV-19/England/CAMC-AAFD3F/2020\|EPI_ISL_623411\|20 | ···················· | ···················· | ···················· | ···················· | ···················· | ···················· |
| P | 724 | hCoV-19/England/MILK-AB8692/2020\|EPI_ISL_623412\|20 | ···················· | ···················· | ···················· | ···················· | ···················· | ···················· |
| P | 725 | hCoV-19/England/MILK-ABB6EA/2020\|EPI_ISL_623413\|20 | ···················· | ·A·················· | ···················· | ···················· | ···················· | ···················· |
| P | 726 | hCoV-19/England/MILK-AB7A5B/2020\|EPI_ISL_623415\|20 | ···················· | ···················· | ···················· | ···················· | ···················· | ···················· |
| P | 727 | hCoV-19/England/MILK-ABB741/2020\|EPI_ISL_623416\|20 | ···················· | ···················· | ···················· | ···················· | ···················· | ···················· |
| P | 728 | hCoV-19/England/MILK-AB7B0D/2020\|EPI_ISL_623419\|20 | ···················· | ···················· | ···················· | ···················· | ···················· | ···················· |
| P | 729 | hCoV-19/England/MILK-AB82CB/2020\|EPI_ISL_623420\|20 | ···················· | ···················· | ···················· | ···················· | ···················· | ···················· |
| P | 730 | hCoV-19/England/MILK-AB7AF1/2020\|EPI_ISL_623422\|20 | ···················· | ···················· | ···················· | ···················· | ···················· | ···················· |
| P | 731 | hCoV-19/England/MILK-AB83AA/2020\|EPI_ISL_623423\|20 | ···················· | ···················· | ···················· | ···················· | ······T············· | ···················· |
| P | 732 | hCoV-19/England/CAMC-AAFC14/2020\|EPI_ISL_623424\|20 | ···················· | ···················· | ···················· | ···················· | ···················· | ···················· |
| P | 733 | hCoV-19/England/CAMC-AAFD7B/2020\|EPI_ISL_623425\|20 | ···················· | ···················· | ···················· | ···················· | ······T············· | ···················· |
| P | 734 | hCoV-19/England/CAMC-AAFA92/2020\|EPI_ISL_623426\|20 | ···················· | ·A·················· | ···················· | ···················· | ···················· | ··A················· |
| P | 735 | hCoV-19/England/ALDP-A9591A/2020\|EPI_ISL_624680\|20 | ···················· | ···················· | ···················· | ···················· | ···················· | ···················· |
| P | 736 | hCoV-19/England/ALDP-A94641/2020\|EPI_ISL_624681\|20 | ···················· | ···················· | ···················· | ···················· | ···················· | ···················· |
| P | 737 | hCoV-19/England/ALDP-A95EC0/2020\|EPI_ISL_624682\|20 | ···················· | ···················· | ···················· | ···················· | ···················· | ···················· |
| P | 738 | hCoV-19/England/ALDP-A95ADB/2020\|EPI_ISL_624683\|20 | ···················· | ···················· | ···················· | ···················· | ···················· | ···················· |
| P | 739 | hCoV-19/England/ALDP-A94AEB/2020\|EPI_ISL_624684\|20 | ···················· | ···················· | ···················· | ···················· | ···················· | ···················· |
| P | 740 | hCoV-19/England/ALDP-A94526/2020\|EPI_ISL_624686\|20 | ···················· | ···················· | ···················· | ···················· | ···················· | ···················· |
| P | 741 | hCoV-19/England/ALDP-A96120/2020\|EPI_ISL_624688\|20 | ···················· | ···················· | ···················· | ···················· | ···················· | ···················· |
| P | 742 | hCoV-19/England/ALDP-A95E2A/2020\|EPI_ISL_624689\|20 | ···················· | ···················· | ···················· | ···················· | ···················· | ···················· |
| P | 743 | hCoV-19/England/ALDP-A9615D/2020\|EPI_ISL_624690\|20 | ···················· | ···················· | ···················· | ···················· | ···················· | ···················· |
| P | 744 | hCoV-19/Wales/ALDP-A955AD/2020\|EPI_ISL_624691\|2020 | ···················· | ···················· | ···················· | ···················· | ···················· | ···················· |
| P | 745 | hCoV-19/England/ALDP-A944CF/2020\|EPI_ISL_624692\|20 | ···················· | ···················· | ···················· | ···················· | ···················· | ···················· |
| P | 746 | hCoV-19/England/ALDP-A95394/2020\|EPI_ISL_624694\|20 | ···················· | ···················· | ···················· | ···················· | ···················· | ···················· |
| P | 747 | hCoV-19/England/ALDP-A946D8/2020\|EPI_ISL_624696\|20 | ···················· | ···················· | ···················· | ···················· | ···················· | ···················· |
| P | 748 | hCoV-19/England/ALDP-A94720/2020\|EPI_ISL_624697\|20 | ···················· | ···················· | ···················· | ···················· | ···················· | ···················· |
| P | 749 | hCoV-19/England/ALDP-A964DC/2020\|EPI_ISL_624698\|20 | ···················· | ···················· | ···················· | ···················· | ···················· | ···················· |
| P | 750 | hCoV-19/England/ALDP-A95E57/2020\|EPI_ISL_624736\|20 | ···················· | ···················· | ···················· | ···················· | ···················· | ···················· |
| P | 751 | hCoV-19/England/ALDP-A94A18/2020\|EPI_ISL_624737\|20 | ···················· | ·A·······A·········· | ···················· | ···················· | ···················· | ···················· |
| P | 752 | hCoV-19/England/ALDP-A9459F/2020\|EPI_ISL_624738\|20 | ···················· | ···················· | ···················· | ···················· | ···················· | ···················· |
| P | 753 | hCoV-19/England/ALDP-A94AFA/2020\|EPI_ISL_624739\|20 | ···················· | ···················· | ···················· | ···················· | ···················· | ···················· |

| | | | | | | | | |
|---|---|---|---|---|---|---|---|---|
| P | 754 | hCoV-19/England/ALDP-A95604/2020\|EPI_ISL_624741\|20 | ···················· | ···················· | ···················· | ···················· | ···················· | ···················· |
| P | 755 | hCoV-19/England/ALDP-A95D3C/2020\|EPI_ISL_624743\|20 | ···················· | ···················· | ···················· | ···················· | ···················· | ···················· |
| P | 756 | hCoV-19/England/ALDP-A95CF3/2020\|EPI_ISL_624744\|20 | ···················· | ···················· | ···················· | ···················· | ···················· | ···················· |
| P | 757 | hCoV-19/England/ALDP-A947A8/2020\|EPI_ISL_624745\|20 | ···················· | ···················· | ···················· | ···················· | ···················· | ···················· |
| P | 758 | hCoV-19/England/ALDP-A95ABD/2020\|EPI_ISL_624747\|20 | ···················· | ···················· | ···················· | ···················· | ···················· | ···················· |
| P | 759 | hCoV-19/England/ALDP-A96032/2020\|EPI_ISL_624748\|20 | ···················· | ···················· | ···················· | ···················· | ···················· | ···················· |
| P | 760 | hCoV-19/England/ALDP-A958FF/2020\|EPI_ISL_624749\|20 | ···················· | ···················· | ···················· | ···················· | ···················· | ···················· |
| P | 761 | hCoV-19/England/ALDP-A95C99/2020\|EPI_ISL_624750\|20 | ···················· | ···················· | ···················· | ···················· | ···················· | ···················· |
| P | 762 | hCoV-19/England/ALDP-A95CE4/2020\|EPI_ISL_624751\|20 | ···················· | ···················· | ···················· | ···················· | ···················· | ···················· |
| P | 763 | hCoV-19/England/ALDP-A964AF/2020\|EPI_ISL_624752\|20 | ···················· | ···················· | ···················· | ···················· | ···················· | ···················· |
| P | 764 | hCoV-19/England/ALDP-A94CF4/2020\|EPI_ISL_624753\|20 | ···················· | ···················· | ···················· | ···················· | ···················· | ···················· |
| P | 765 | hCoV-19/Wales/ALDP-A95F63/2020\|EPI_ISL_624754\|2020 | ···················· | ···················· | ···················· | ···················· | ···················· | ···················· |
| P | 766 | hCoV-19/England/ALDP-A96357/2020\|EPI_ISL_624755\|20 | ···················· | ·A·················· | ···················· | ···················· | ···················· | ···················· |
| P | 767 | hCoV-19/England/ALDP-A960C9/2020\|EPI_ISL_624756\|20 | ···················· | ···················· | ···················· | ···················· | ···················· | ···················· |
| P | 768 | hCoV-19/England/ALDP-A94483/2020\|EPI_ISL_624757\|20 | ···················· | ···················· | ···················· | ···················· | ···················· | ···················· |
| P | 769 | hCoV-19/Denmark/DCGC-3381/2020\|EPI_ISL_616140\|2020 | ···················· | ···················· | ···················· | ···················· | ···················· | ···················· |
| P | 770 | hCoV-19/Denmark/DCGC-3382/2020\|EPI_ISL_616141\|2020 | ···················· | ···················· | ···················· | ···················· | ···················· | ···················· |
| P | 771 | hCoV-19/Denmark/DCGC-3383/2020\|EPI_ISL_616142\|2020 | ···················· | ···················· | ···················· | ···················· | ···················· | ···················· |
| P | 772 | hCoV-19/Denmark/DCGC-3384/2020\|EPI_ISL_616143\|2020 | ···················· | ···················· | ···················· | ···················· | ···················· | ···················· |
| P | 773 | hCoV-19/Denmark/DCGC-3385/2020\|EPI_ISL_616144\|2020 | ···················· | ···················· | ···················· | ···················· | ···················· | ···················· |
| P | 774 | hCoV-19/Denmark/DCGC-3386/2020\|EPI_ISL_616145\|2020 | ···················· | ···················· | ···················· | ···················· | ···················· | ···················· |
| P | 775 | hCoV-19/Denmark/DCGC-3387/2020\|EPI_ISL_616146\|2020 | ···················· | ···················· | ···················· | ···················· | ···················· | ···················· |
| P | 776 | hCoV-19/Denmark/DCGC-3388/2020\|EPI_ISL_616147\|2020 | ···················· | ···················· | ···················· | ···················· | ···················· | ···················· |
| P | 777 | hCoV-19/Denmark/DCGC-3389/2020\|EPI_ISL_616148\|2020 | ···················· | ···················· | ···················· | ···················· | ···················· | ···················· |
| P | 778 | hCoV-19/Denmark/DCGC-3390/2020\|EPI_ISL_616149\|2020 | ···················· | ···················· | ···················· | ···················· | ···················· | ···················· |
| P | 779 | hCoV-19/Denmark/DCGC-3391/2020\|EPI_ISL_616150\|2020 | ···················· | ·A·················· | ···················· | ···················· | ···················· | ··A················· |
| P | 780 | hCoV-19/Denmark/DCGC-3392/2020\|EPI_ISL_616151\|2020 | ···················· | ···················· | ···················· | ···················· | ···················· | ···················· |
| P | 781 | hCoV-19/Denmark/DCGC-3393/2020\|EPI_ISL_616152\|2020 | ···················· | ···················· | ···················· | ···················· | ···················· | ···················· |
| P | 782 | hCoV-19/Denmark/DCGC-3394/2020\|EPI_ISL_616153\|2020 | ···················· | ···················· | ···················· | ···················· | ···················· | ···················· |
| P | 783 | hCoV-19/Denmark/DCGC-3395/2020\|EPI_ISL_616154\|2020 | ···················· | ···················· | ···················· | ···················· | ···················· | ···················· |
| P | 784 | hCoV-19/Denmark/DCGC-3396/2020\|EPI_ISL_616155\|2020 | ···················· | ···················· | ···················· | ···················· | ···················· | ···················· |
| P | 785 | hCoV-19/Denmark/DCGC-3397/2020\|EPI_ISL_616156\|2020 | ···················· | ···················· | ···················· | ···················· | ···················· | ···················· |
| P | 786 | hCoV-19/Denmark/DCGC-3398/2020\|EPI_ISL_616157\|2020 | ···················· | ···················· | ···················· | ···················· | ···················· | ···················· |
| P | 787 | hCoV-19/Denmark/DCGC-3399/2020\|EPI_ISL_616158\|2020 | ···················· | ···················· | ···················· | ···················· | ···················· | ···················· |
| P | 788 | hCoV-19/Denmark/DCGC-3400/2020\|EPI_ISL_616159\|2020 | ···················· | ···················· | ···················· | ···················· | ···················· | ···················· |
| P | 789 | hCoV-19/Denmark/DCGC-3401/2020\|EPI_ISL_616160\|2020 | ···················· | ···················· | ···················· | ···················· | ···················· | ···················· |
| P | 790 | hCoV-19/Denmark/DCGC-3402/2020\|EPI_ISL_616161\|2020 | ···················· | ···················· | ···················· | ···················· | ···················· | ···················· |
| P | 791 | hCoV-19/Denmark/DCGC-3403/2020\|EPI_ISL_616162\|2020 | ···················· | ···················· | ···················· | ···················· | ···················· | ···················· |
| P | 792 | hCoV-19/Denmark/DCGC-3404/2020\|EPI_ISL_616163\|2020 | ···················· | ···················· | ···················· | ···················· | ···················· | ···················· |
| P | 793 | hCoV-19/Denmark/DCGC-3405/2020\|EPI_ISL_616164\|2020 | ···················· | ···················· | ···················· | ···················· | ···················· | ···················· |
| P | 794 | hCoV-19/Denmark/DCGC-3406/2020\|EPI_ISL_616165\|2020 | ···················· | ···················· | ···················· | ···················· | ···················· | ···················· |
| P | 795 | hCoV-19/Denmark/DCGC-3407/2020\|EPI_ISL_616166\|2020 | ···················· | ···················· | ···················· | ···················· | ···················· | ···················· |
| P | 796 | hCoV-19/Denmark/DCGC-3408/2020\|EPI_ISL_616167\|2020 | ···················· | ···················· | ···················· | ···················· | ···················· | ···················· |
| P | 797 | hCoV-19/Denmark/DCGC-3409/2020\|EPI_ISL_616168\|2020 | ···················· | ···················· | ···················· | ···················· | ···················· | ···················· |
| P | 798 | hCoV-19/Denmark/DCGC-3410/2020\|EPI_ISL_616169\|2020 | ···················· | ···················· | ···················· | ···················· | ···················· | ···················· |
| P | 799 | hCoV-19/Denmark/DCGC-3411/2020\|EPI_ISL_616170\|2020 | ···················· | ···················· | ···················· | ···················· | ···················· | ···················· |
| P | 800 | hCoV-19/Denmark/DCGC-3412/2020\|EPI_ISL_616171\|2020 | ···················· | ···················· | ···················· | ···················· | ···················· | ···················· |
| P | 801 | hCoV-19/Denmark/DCGC-3413/2020\|EPI_ISL_616172\|2020 | ···················· | ···················· | ···················· | ···················· | ···················· | ···················· |
| P | 802 | hCoV-19/Denmark/DCGC-3414/2020\|EPI_ISL_616173\|2020 | ···················· | ···················· | ···················· | ···················· | ···················· | ···················· |
| P | 803 | hCoV-19/Denmark/DCGC-3415/2020\|EPI_ISL_616174\|2020 | ···················· | ···················· | ···················· | ···················· | ···················· | ···················· |
| P | 804 | hCoV-19/Denmark/DCGC-3417/2020\|EPI_ISL_616176\|2020 | ···················· | ·A·················· | ···················· | ···················· | ···················· | ···················· |
| P | 805 | hCoV-19/Denmark/DCGC-3418/2020\|EPI_ISL_616177\|2020 | ···················· | ···················· | ···················· | ···················· | ···················· | ···················· |
| P | 806 | hCoV-19/Denmark/DCGC-3419/2020\|EPI_ISL_616178\|2020 | ···················· | ···················· | ···················· | ···················· | ···················· | ···················· |
| P | 807 | hCoV-19/Denmark/DCGC-3420/2020\|EPI_ISL_616179\|2020 | ···················· | ···················· | ···················· | ···················· | ···················· | ···················· |
| P | 808 | hCoV-19/Denmark/DCGC-3421/2020\|EPI_ISL_616180\|2020 | ···················· | ···················· | ···················· | ···················· | ···················· | ···················· |
| P | 809 | hCoV-19/Denmark/DCGC-3264/2020\|EPI_ISL_616182\|2020 | ···················· | ···················· | ···················· | ···················· | ···················· | ···················· |
| P | 810 | hCoV-19/Denmark/DCGC-3265/2020\|EPI_ISL_616183\|2020 | ···················· | ·A·················· | ···················· | ···················· | ···················· | ··A················· |
| P | 811 | hCoV-19/Denmark/DCGC-3266/2020\|EPI_ISL_616184\|2020 | ···················· | ···················· | ···················· | ···················· | ···················· | ···················· |
| P | 812 | hCoV-19/Denmark/DCGC-3423/2020\|EPI_ISL_616185\|2020 | ···················· | ···················· | ···················· | ···················· | ···················· | ···················· |
| P | 813 | hCoV-19/Denmark/DCGC-3424/2020\|EPI_ISL_616186\|2020 | ···················· | ···················· | ···················· | ···················· | ···················· | ···················· |
| P | 814 | hCoV-19/Denmark/DCGC-3425/2020\|EPI_ISL_616187\|2020 | ···················· | ···················· | ···················· | ···················· | ···················· | ···················· |
| P | 815 | hCoV-19/Denmark/DCGC-3426/2020\|EPI_ISL_616188\|2020 | ···················· | ···················· | ···················· | ···················· | ···················· | ···················· |
| P | 816 | hCoV-19/Denmark/DCGC-3427/2020\|EPI_ISL_616189\|2020 | ···················· | ···················· | ···················· | ···················· | ···················· | ···················· |
| P | 817 | hCoV-19/Denmark/DCGC-3430/2020\|EPI_ISL_616192\|2020 | ···················· | ···················· | ···················· | ···················· | ···················· | ···················· |
| P | 818 | hCoV-19/Denmark/DCGC-3432/2020\|EPI_ISL_616194\|2020 | ···················· | ···················· | ···················· | ···················· | ···················· | ···················· |
| P | 819 | hCoV-19/Denmark/DCGC-3433/2020\|EPI_ISL_616195\|2020 | ···················· | ···················· | ···················· | ···················· | ···················· | ···················· |
| P | 820 | hCoV-19/Denmark/DCGC-3434/2020\|EPI_ISL_616196\|2020 | ···················· | ···················· | ···················· | ···················· | ···················· | ···················· |
| P | 821 | hCoV-19/Denmark/DCGC-3435/2020\|EPI_ISL_616197\|2020 | ···················· | ···················· | ···················· | ···················· | ···················· | ···················· |
| P | 822 | hCoV-19/Denmark/DCGC-3436/2020\|EPI_ISL_616198\|2020 | ···················· | ···················· | ···················· | ···················· | ···················· | ···················· |
| P | 823 | hCoV-19/Denmark/DCGC-3437/2020\|EPI_ISL_616199\|2020 | ···················· | ···················· | ···················· | ···················· | ···················· | ···················· |
| P | 824 | hCoV-19/Denmark/DCGC-3438/2020\|EPI_ISL_616200\|2020 | ···················· | ···················· | ···················· | ···················· | ···················· | ···················· |
| P | 825 | hCoV-19/Denmark/DCGC-3439/2020\|EPI_ISL_616201\|2020 | ···················· | ···················· | ···················· | ···················· | ···················· | ···················· |
| P | 826 | hCoV-19/Denmark/DCGC-3441/2020\|EPI_ISL_616203\|2020 | ···················· | ···················· | ···················· | ···················· | ···················· | ···················· |
| P | 827 | hCoV-19/Denmark/DCGC-3442/2020\|EPI_ISL_616204\|2020 | ···················· | ···················· | ···················· | ···················· | ···················· | ···················· |
| P | 828 | hCoV-19/Denmark/DCGC-3443/2020\|EPI_ISL_616205\|2020 | ···················· | ···················· | ···················· | ···················· | ···················· | ···················· |
| P | 829 | hCoV-19/Denmark/DCGC-3444/2020\|EPI_ISL_616206\|2020 | ···················· | ···················· | ···················· | ···················· | ···················· | ···················· |
| P | 830 | hCoV-19/Denmark/DCGC-3445/2020\|EPI_ISL_616207\|2020 | ···················· | ···················· | ···················· | ···················· | ···················· | ···················· |
| P | 831 | hCoV-19/Denmark/DCGC-3446/2020\|EPI_ISL_616208\|2020 | ···················· | ···················· | ···················· | ···················· | ···················· | ···················· |
| P | 832 | hCoV-19/Denmark/DCGC-3267/2020\|EPI_ISL_616209\|2020 | ···················· | ···················· | ···················· | ···················· | ···················· | ···················· |
| P | 833 | hCoV-19/Denmark/DCGC-3447/2020\|EPI_ISL_616210\|2020 | ···················· | ···················· | ···················· | ···················· | ···················· | ···················· |

| | | | | | | | |
|---|---|---|---|---|---|---|---|
| P | 834 hCoV-19/Denmark/DCGC-3448/2020\|EPI_ISL_616211\|2020 | ···················· | ···················· | ····················· | ···················· | ···················· | ···················· |
| P | 835 hCoV-19/Denmark/DCGC-3268/2020\|EPI_ISL_616212\|2020 | ···················· | ···················· | ····················· | ···················· | ···················· | ···················· |
| P | 836 hCoV-19/Denmark/DCGC-3449/2020\|EPI_ISL_616213\|2020 | ···················· | ···················· | ····················· | ···················· | ···················· | ···················· |
| P | 837 hCoV-19/Denmark/DCGC-3450/2020\|EPI_ISL_616214\|2020 | ···················· | ···················· | ····················· | ···················· | ···················· | ···················· |
| P | 838 hCoV-19/Denmark/DCGC-3451/2020\|EPI_ISL_616215\|2020 | ···················· | ···················· | ····················· | ···················· | ···················· | ···················· |
| P | 839 hCoV-19/Denmark/DCGC-3452/2020\|EPI_ISL_616216\|2020 | ···················· | ···················· | ····················· | ···················· | ···················· | ···················· |
| P | 840 hCoV-19/Denmark/DCGC-3453/2020\|EPI_ISL_616217\|2020 | ···················· | ···················· | ····················· | ···················· | ···················· | ···················· |
| P | 841 hCoV-19/Denmark/DCGC-3454/2020\|EPI_ISL_616218\|2020 | ···················· | ···················· | ····················· | ···················· | ···················· | ···················· |
| P | 842 hCoV-19/Denmark/DCGC-3455/2020\|EPI_ISL_616219\|2020 | ···················· | ···················· | ····················· | ···················· | ···················· | ···················· |
| P | 843 hCoV-19/Denmark/DCGC-3589/2020\|EPI_ISL_616232\|2020 | ···················· | ···················· | ····················· | ···················· | ···················· | ···················· |
| P | 844 hCoV-19/Denmark/DCGC-3590/2020\|EPI_ISL_616233\|2020 | ···················· | ···················· | ····················· | ···················· | ···················· | ···················· |
| P | 845 hCoV-19/Denmark/DCGC-3591/2020\|EPI_ISL_616234\|2020 | ···················· | ···················· | ····················· | ···················· | ···················· | ···················· |
| P | 846 hCoV-19/Denmark/DCGC-3592/2020\|EPI_ISL_616235\|2020 | ···················· | ···················· | ····················· | ···················· | ···················· | ···················· |
| P | 847 hCoV-19/Denmark/DCGC-3597/2020\|EPI_ISL_616240\|2020 | ···················· | ···················· | ····················· | ···················· | ···················· | ···················· |
| P | 848 hCoV-19/Denmark/DCGC-3598/2020\|EPI_ISL_616241\|2020 | ········T··········· | ···················· | ····················· | ···················· | ···················· | ···················· |
| P | 849 hCoV-19/Denmark/DCGC-3599/2020\|EPI_ISL_616242\|2020 | ···················· | ···················· | ····················· | ···················· | ···················· | ···················· |
| P | 850 hCoV-19/Denmark/DCGC-3600/2020\|EPI_ISL_616243\|2020 | ···················· | ···················· | ····················· | ···················· | ···················· | ···················· |
| P | 851 hCoV-19/Denmark/DCGC-3601/2020\|EPI_ISL_616244\|2020 | ···················· | ···················· | ····················· | ···················· | ···················· | ···················· |
| P | 852 hCoV-19/Denmark/DCGC-3602/2020\|EPI_ISL_616245\|2020 | ···················· | ···················· | ····················· | ···················· | ···················· | ···················· |
| P | 853 hCoV-19/Denmark/DCGC-3603/2020\|EPI_ISL_616246\|2020 | ···················· | ···················· | ····················· | ···················· | ···················· | ···················· |
| P | 854 hCoV-19/Denmark/DCGC-3604/2020\|EPI_ISL_616247\|2020 | ···················· | ···················· | ····················· | ···················· | ···················· | ···················· |
| P | 855 hCoV-19/Denmark/DCGC-3461/2020\|EPI_ISL_616248\|2020 | ···················· | ···················· | ····················· | ···················· | ···················· | ···················· |
| P | 856 hCoV-19/Denmark/DCGC-3462/2020\|EPI_ISL_616249\|2020 | ···················· | ·A·················· | ····················· | ···················· | ···················· | ···A················ |
| P | 857 hCoV-19/Denmark/DCGC-3463/2020\|EPI_ISL_616250\|2020 | ···················· | ···················· | ····················· | ···················· | ···················· | ···················· |
| P | 858 hCoV-19/Denmark/DCGC-3464/2020\|EPI_ISL_616251\|2020 | ···················· | ···················· | ····················· | ···················· | ···················· | ···················· |
| P | 859 hCoV-19/Denmark/DCGC-3466/2020\|EPI_ISL_616253\|2020 | ···················· | ···················· | ····················· | ···················· | ···················· | ···················· |
| P | 860 hCoV-19/Denmark/DCGC-3467/2020\|EPI_ISL_616254\|2020 | ···················· | ···················· | ····················· | ···················· | ···················· | ···················· |
| P | 861 hCoV-19/Denmark/DCGC-3468/2020\|EPI_ISL_616255\|2020 | ···················· | ···················· | ····················· | ···················· | ···················· | ···················· |
| P | 862 hCoV-19/Denmark/DCGC-3469/2020\|EPI_ISL_616256\|2020 | ···················· | ···················· | ····················· | ···················· | ···················· | ···················· |
| P | 863 hCoV-19/Denmark/DCGC-3470/2020\|EPI_ISL_616257\|2020 | ···················· | ···················· | ····················· | ···················· | ···················· | ···················· |
| P | 864 hCoV-19/Denmark/DCGC-3471/2020\|EPI_ISL_616258\|2020 | ···················· | ···················· | ····················· | ···················· | ···················· | ···················· |
| P | 865 hCoV-19/Denmark/DCGC-3472/2020\|EPI_ISL_616259\|2020 | ···················· | ···················· | ····················· | ···················· | ···················· | ···················· |
| P | 866 hCoV-19/Denmark/DCGC-2171/2020\|EPI_ISL_617470\|2020 | ···················· | ···················· | ····················· | ···················· | ···················· | ···················· |
| P | 867 hCoV-19/Denmark/DCGC-2172/2020\|EPI_ISL_617471\|2020 | ···················· | ···················· | ····················· | ···················· | ···················· | ···················· |
| P | 868 hCoV-19/Denmark/DCGC-2173/2020\|EPI_ISL_617472\|2020 | ···················· | ···················· | ····················· | ···················· | ···················· | ···················· |
| P | 869 hCoV-19/Denmark/DCGC-2174/2020\|EPI_ISL_617473\|2020 | ···················· | ···················· | ····················· | ···················· | ···················· | ···················· |
| P | 870 hCoV-19/Denmark/DCGC-2176/2020\|EPI_ISL_617475\|2020 | ···················· | ···················· | ····················· | ···················· | ···················· | ···················· |
| P | 871 hCoV-19/Denmark/DCGC-2177/2020\|EPI_ISL_617476\|2020 | ···················· | ···················· | ····················· | ···················· | ···················· | ···················· |
| P | 872 hCoV-19/Denmark/DCGC-2178/2020\|EPI_ISL_617477\|2020 | ···················· | ···················· | ····················· | ···················· | ···················· | ···················· |
| P | 873 hCoV-19/Denmark/DCGC-2179/2020\|EPI_ISL_617478\|2020 | ···················· | ···················· | ····················· | ···················· | ···················· | ···················· |
| P | 874 hCoV-19/Denmark/DCGC-2180/2020\|EPI_ISL_617479\|2020 | ···················· | ···················· | ····················· | ···················· | ···················· | ···················· |
| P | 875 hCoV-19/Denmark/DCGC-2181/2020\|EPI_ISL_617480\|2020 | ···················· | ···················· | ····················· | ···················· | ···················· | ···················· |
| P | 876 hCoV-19/Denmark/DCGC-2182/2020\|EPI_ISL_617481\|2020 | ···················· | ···················· | ····················· | ···················· | ···················· | ···················· |
| P | 877 hCoV-19/Denmark/DCGC-2183/2020\|EPI_ISL_617482\|2020 | ···················· | ···················· | ····················· | ···················· | ···················· | ···················· |
| P | 878 hCoV-19/Denmark/DCGC-2184/2020\|EPI_ISL_617483\|2020 | ···················· | ···················· | ····················· | ···················· | ···················· | ···················· |
| P | 879 hCoV-19/Denmark/DCGC-2423/2020\|EPI_ISL_617484\|2020 | ···················· | ···················· | ····················· | ···················· | ···················· | ···················· |
| P | 880 hCoV-19/Denmark/DCGC-2185/2020\|EPI_ISL_617485\|2020 | ···················· | ···················· | ····················· | ···················· | ···················· | ···················· |
| P | 881 hCoV-19/Denmark/DCGC-2186/2020\|EPI_ISL_617486\|2020 | ···················· | ···················· | ····················· | ···················· | ···················· | ···················· |
| P | 882 hCoV-19/Denmark/DCGC-2424/2020\|EPI_ISL_617487\|2020 | ···················· | ···················· | ····················· | ···················· | ···················· | ···················· |
| P | 883 hCoV-19/Denmark/DCGC-2187/2020\|EPI_ISL_617488\|2020 | ···················· | ···················· | ····················· | ···················· | ···················· | ···················· |
| P | 884 hCoV-19/Denmark/DCGC-2188/2020\|EPI_ISL_617489\|2020 | ···················· | ···G················ | ····················· | ···················· | ···················· | ···················· |
| P | 885 hCoV-19/Denmark/DCGC-2189/2020\|EPI_ISL_617490\|2020 | ···················· | ·A·················· | ····················· | ···················· | ···················· | ···················· |
| P | 886 hCoV-19/Denmark/DCGC-2191/2020\|EPI_ISL_617492\|2020 | ···················· | ···················· | ····················· | ···················· | ···················· | ···················· |
| P | 887 hCoV-19/Denmark/DCGC-2192/2020\|EPI_ISL_617493\|2020 | ···················· | ···················· | ····················· | ···················· | ···················· | ···················· |
| P | 888 hCoV-19/Denmark/DCGC-2193/2020\|EPI_ISL_617494\|2020 | ···················· | ···················· | ····················· | ···················· | ···················· | ···················· |
| P | 889 hCoV-19/Denmark/DCGC-2194/2020\|EPI_ISL_617495\|2020 | ···················· | ···················· | ····················· | ···················· | ···················· | ···················· |
| P | 890 hCoV-19/Denmark/DCGC-2195/2020\|EPI_ISL_617496\|2020 | ···················· | ···················· | ····················· | ···················· | ···················· | ···················· |
| P | 891 hCoV-19/Denmark/DCGC-2197/2020\|EPI_ISL_617498\|2020 | ···················· | ···················· | ····················· | ···················· | ···················· | ···················· |
| P | 892 hCoV-19/Denmark/DCGC-2198/2020\|EPI_ISL_617499\|2020 | ···················· | ···················· | ····················· | ···················· | ···················· | ···················· |
| P | 893 hCoV-19/Denmark/DCGC-2206/2020\|EPI_ISL_617509\|2020 | ···················· | ···················· | ····················· | ···················· | ···················· | ···················· |
| P | 894 hCoV-19/Denmark/DCGC-2207/2020\|EPI_ISL_617510\|2020 | ···················· | ···················· | ····················· | ···················· | ···················· | ···················· |
| P | 895 hCoV-19/Denmark/DCGC-2208/2020\|EPI_ISL_617511\|2020 | ···················· | ···················· | ····················· | ···················· | ···················· | ···················· |
| P | 896 hCoV-19/Denmark/DCGC-2209/2020\|EPI_ISL_617512\|2020 | ···················· | ···················· | ····················· | ···················· | ···················· | ···················· |
| P | 897 hCoV-19/Denmark/DCGC-2210/2020\|EPI_ISL_617513\|2020 | ···················· | ···················· | ····················· | ···················· | ···················· | ···················· |
| P | 898 hCoV-19/Denmark/DCGC-2211/2020\|EPI_ISL_617514\|2020 | ···················· | ···················· | ····················· | ···················· | ···················· | ···················· |
| P | 899 hCoV-19/Denmark/DCGC-2212/2020\|EPI_ISL_617515\|2020 | ···················· | ···················· | ····················· | ···················· | ···················· | ···················· |
| P | 900 hCoV-19/Denmark/DCGC-2213/2020\|EPI_ISL_617516\|2020 | ···················· | ···················· | ····················· | ···················· | ···················· | ···················· |
| P | 901 hCoV-19/Denmark/DCGC-2214/2020\|EPI_ISL_617517\|2020 | ···················· | ···················· | ····················· | ···················· | ···················· | ···················· |
| P | 902 hCoV-19/Denmark/DCGC-2215/2020\|EPI_ISL_617518\|2020 | ···················· | ···················· | ····················· | ···················· | ···················· | ···················· |
| P | 903 hCoV-19/Denmark/DCGC-2216/2020\|EPI_ISL_617519\|2020 | ···················· | ···················· | ····················· | ···················· | ···················· | ···················· |
| P | 904 hCoV-19/Denmark/DCGC-2217/2020\|EPI_ISL_617520\|2020 | ···················· | ···················· | ····················· | ···················· | ···················· | ···················· |
| P | 905 hCoV-19/Denmark/DCGC-2218/2020\|EPI_ISL_617521\|2020 | ···················· | ···················· | ····················· | ···················· | ···················· | ···················· |
| P | 906 hCoV-19/Denmark/DCGC-2427/2020\|EPI_ISL_617522\|2020 | ···················· | ···················· | ····················· | ···················· | ···················· | ···················· |
| P | 907 hCoV-19/Denmark/DCGC-2428/2020\|EPI_ISL_617524\|2020 | ···················· | ···················· | ····················· | ···················· | ···················· | ···················· |
| P | 908 hCoV-19/Denmark/DCGC-2220/2020\|EPI_ISL_617525\|2020 | ···················· | ···················· | ····················· | ···················· | ···················· | ···················· |
| P | 909 hCoV-19/Denmark/DCGC-2221/2020\|EPI_ISL_617526\|2020 | ···················· | ···················· | ····················· | ···················· | ···················· | ···················· |
| P | 910 hCoV-19/Denmark/DCGC-2222/2020\|EPI_ISL_617527\|2020 | ···················· | ···················· | ····················· | ···················· | ···················· | ···················· |
| P | 911 hCoV-19/Denmark/DCGC-2223/2020\|EPI_ISL_617528\|2020 | ···················· | ···················· | ····················· | ···················· | ···················· | ···················· |
| P | 912 hCoV-19/Denmark/DCGC-2225/2020\|EPI_ISL_617530\|2020 | ···················· | ···················· | ····················· | ···················· | ···················· | ···················· |
| P | 913 hCoV-19/Denmark/DCGC-2226/2020\|EPI_ISL_617531\|2020 | ···················· | ···················· | ····················· | ···················· | ···················· | ···················· |

| | | | | | | | | |
|---|---|---|---|---|---|---|---|---|
| P | 914 | hCoV-19/Denmark/DCGC-2227/2020\|EPI_ISL_617532\|2020 | ···················· | ···················· | ···················· | ···················· | ···················· | ···················· |
| P | 915 | hCoV-19/Denmark/DCGC-2228/2020\|EPI_ISL_617533\|2020 | ···················· | ···················· | ···················· | ···················· | ···················· | ···················· |
| P | 916 | hCoV-19/Denmark/DCGC-2229/2020\|EPI_ISL_617534\|2020 | ···················· | ···················· | ···················· | ···················· | ···················· | ···················· |
| P | 917 | hCoV-19/Denmark/DCGC-2230/2020\|EPI_ISL_617535\|2020 | ···················· | ···················· | ···················· | ···················· | ···················· | ···················· |
| P | 918 | hCoV-19/Denmark/DCGC-2232/2020\|EPI_ISL_617537\|2020 | ···················· | ·A·················· | ···················· | ···················· | ···················· | ···················· |
| P | 919 | hCoV-19/Denmark/DCGC-2233/2020\|EPI_ISL_617538\|2020 | ···················· | ···················· | ···················· | ···················· | ···················· | ···················· |
| P | 920 | hCoV-19/Denmark/DCGC-2234/2020\|EPI_ISL_617539\|2020 | ···················· | ···················· | ···················· | ···················· | ···················· | ···················· |
| P | 921 | hCoV-19/Denmark/DCGC-2236/2020\|EPI_ISL_617541\|2020 | ···················· | ···················· | ···················· | ···················· | ···················· | ···················· |
| P | 922 | hCoV-19/Denmark/DCGC-2237/2020\|EPI_ISL_617542\|2020 | ···················· | ···················· | ···················· | ···················· | ···················· | ···················· |
| P | 923 | hCoV-19/Denmark/DCGC-2238/2020\|EPI_ISL_617543\|2020 | ···················· | ···················· | ···················· | ···················· | ···················· | ···················· |
| P | 924 | hCoV-19/Denmark/DCGC-2239/2020\|EPI_ISL_617544\|2020 | ···················· | ···················· | ···················· | ···················· | ···················· | ···················· |
| P | 925 | hCoV-19/Denmark/DCGC-2240/2020\|EPI_ISL_617545\|2020 | ···················· | ···················· | ···················· | ···················· | ···················· | ···················· |
| P | 926 | hCoV-19/Denmark/DCGC-2241/2020\|EPI_ISL_617546\|2020 | ···················· | ···················· | ···················· | ···················· | ···················· | ···················· |
| P | 927 | hCoV-19/Denmark/DCGC-2242/2020\|EPI_ISL_617547\|2020 | ···················· | ·A·················· | ···················· | ···················· | ···················· | ···················· |
| P | 928 | hCoV-19/Denmark/DCGC-2243/2020\|EPI_ISL_617548\|2020 | ···················· | ···················· | ···················· | ···················· | ···················· | ···················· |
| P | 929 | hCoV-19/Denmark/DCGC-2244/2020\|EPI_ISL_617549\|2020 | ···················· | ···················· | ···················· | ···················· | ···················· | ···················· |
| P | 930 | hCoV-19/Denmark/DCGC-2380/2020\|EPI_ISL_617560\|2020 | ···················· | ·A··A··············· | ···················· | ···················· | ···················· | ···················· |
| P | 931 | hCoV-19/Denmark/DCGC-2381/2020\|EPI_ISL_617561\|2020 | ···················· | ····G··············· | ···················· | ···················· | ···················· | ···················· |
| P | 932 | hCoV-19/Denmark/DCGC-2382/2020\|EPI_ISL_617562\|2020 | ···················· | ···················· | ···················· | ···················· | ···················· | ···················· |
| P | 933 | hCoV-19/Denmark/DCGC-2469/2020\|EPI_ISL_617570\|2020 | ···················· | ···················· | ···················· | ···················· | ···················· | ···················· |
| P | 934 | hCoV-19/Denmark/DCGC-2390/2020\|EPI_ISL_617571\|2020 | ···················· | ···················· | ···················· | ···················· | ···················· | ···················· |
| P | 935 | hCoV-19/Denmark/DCGC-2391/2020\|EPI_ISL_617572\|2020 | ···················· | ···················· | ···················· | ···················· | ···················· | ···················· |
| P | 936 | hCoV-19/Denmark/DCGC-2392/2020\|EPI_ISL_617574\|2020 | ···················· | ···················· | ···················· | ···················· | ···················· | ···················· |
| P | 937 | hCoV-19/Denmark/DCGC-2393/2020\|EPI_ISL_617575\|2020 | ···················· | ···················· | ···················· | ···················· | ···················· | ···················· |
| P | 938 | hCoV-19/Denmark/DCGC-2395/2020\|EPI_ISL_617577\|2020 | ···················· | ···················· | ···················· | ···················· | ···················· | ···················· |
| P | 939 | hCoV-19/Denmark/DCGC-2397/2020\|EPI_ISL_617579\|2020 | ···················· | ···················· | ···················· | ···················· | ···················· | ···················· |
| P | 940 | hCoV-19/Denmark/DCGC-2399/2020\|EPI_ISL_617581\|2020 | ···················· | ···················· | ···················· | ···················· | ···················· | ···················· |
| P | 941 | hCoV-19/Denmark/DCGC-2400/2020\|EPI_ISL_617582\|2020 | ···················· | ···················· | ···················· | ···················· | ···················· | ···················· |
| P | 942 | hCoV-19/Denmark/DCGC-2401/2020\|EPI_ISL_617583\|2020 | ···················· | ···················· | ···················· | ···················· | ···················· | ···················· |
| P | 943 | hCoV-19/Denmark/DCGC-2402/2020\|EPI_ISL_617584\|2020 | ···················· | ···················· | ···················· | ···················· | ···················· | ···················· |
| P | 944 | hCoV-19/Denmark/DCGC-2403/2020\|EPI_ISL_617585\|2020 | ···················· | ···················· | ···················· | ···················· | ···················· | ···················· |
| P | 945 | hCoV-19/Denmark/DCGC-2404/2020\|EPI_ISL_617586\|2020 | ···················· | ···················· | ···················· | ···················· | ···················· | ···················· |
| P | 946 | hCoV-19/Denmark/DCGC-2405/2020\|EPI_ISL_617587\|2020 | ···················· | ···················· | ···················· | ···················· | ···················· | ···················· |
| P | 947 | hCoV-19/Denmark/DCGC-2406/2020\|EPI_ISL_617588\|2020 | ···················· | ···················· | ···················· | ···················· | ···················· | ···················· |
| P | 948 | hCoV-19/Denmark/DCGC-2407/2020\|EPI_ISL_617589\|2020 | ···················· | ···················· | ···················· | ···················· | ···················· | ···················· |
| P | 949 | hCoV-19/Denmark/DCGC-7901/2020\|EPI_ISL_618840\|2020 | ···················· | ·A·················· | ···················· | ···················· | ···················· | ···················· |
| P | 950 | hCoV-19/Denmark/DCGC-7914/2020\|EPI_ISL_618842\|2020 | ···················· | ···················· | ···················· | ···················· | ···················· | ···················· |
| P | 951 | hCoV-19/Denmark/DCGC-7920/2020\|EPI_ISL_618843\|2020 | ···················· | ···················· | ···················· | ···················· | ···················· | ···················· |
| P | 952 | hCoV-19/Denmark/DCGC-7860/2020\|EPI_ISL_618844\|2020 | ···················· | ···················· | ···················· | ···················· | ···················· | ···················· |
| P | 953 | hCoV-19/Denmark/DCGC-7867/2020\|EPI_ISL_618845\|2020 | ···················· | ···················· | ···················· | ···················· | ···················· | ···················· |
| P | 954 | hCoV-19/Denmark/DCGC-7874/2020\|EPI_ISL_618846\|2020 | ···················· | ···················· | ···················· | ···················· | ···················· | ···················· |
| P | 955 | hCoV-19/Denmark/DCGC-7879/2020\|EPI_ISL_618847\|2020 | ···················· | ·A·················· | ···················· | ···················· | ···················· | ···················· |
| P | 956 | hCoV-19/Denmark/DCGC-7902/2020\|EPI_ISL_618849\|2020 | ···················· | ···················· | ···················· | ···················· | ···················· | ···················· |
| P | 957 | hCoV-19/Denmark/DCGC-7908/2020\|EPI_ISL_618850\|2020 | ···················· | ···················· | ···················· | ···················· | ···················· | ···················· |
| P | 958 | hCoV-19/Denmark/DCGC-7915/2020\|EPI_ISL_618851\|2020 | ···················· | ···················· | ···················· | ···················· | ···················· | ···················· |
| P | 959 | hCoV-19/Denmark/DCGC-7861/2020\|EPI_ISL_618852\|2020 | ···················· | ···················· | ···················· | ···················· | ···················· | ···················· |
| P | 960 | hCoV-19/Denmark/DCGC-7868/2020\|EPI_ISL_618853\|2020 | ···················· | ···················· | ···················· | ···················· | ···················· | ···················· |
| P | 961 | hCoV-19/Denmark/DCGC-7887/2020\|EPI_ISL_618854\|2020 | ···················· | ···················· | ···················· | ···················· | ···················· | ···················· |
| P | 962 | hCoV-19/Denmark/DCGC-7897/2020\|EPI_ISL_618855\|2020 | ···················· | ·A·················· | ···················· | ···················· | ···················· | ···················· |
| P | 963 | hCoV-19/Denmark/DCGC-7909/2020\|EPI_ISL_618856\|2020 | ···················· | ···················· | ···················· | ···················· | ···················· | ···················· |
| P | 964 | hCoV-19/Denmark/DCGC-7855/2020\|EPI_ISL_618857\|2020 | ···················· | ···················· | ···················· | ···················· | ···················· | ···················· |
| P | 965 | hCoV-19/Denmark/DCGC-7869/2020\|EPI_ISL_618858\|2020 | ···················· | ···················· | ···················· | ···················· | ···················· | ···················· |
| P | 966 | hCoV-19/Denmark/DCGC-7875/2020\|EPI_ISL_618859\|2020 | ···················· | ···················· | ···················· | ···················· | ···················· | ···················· |
| P | 967 | hCoV-19/Denmark/DCGC-7880/2020\|EPI_ISL_618860\|2020 | ···················· | ···················· | ···················· | ···················· | ···················· | ···················· |
| P | 968 | hCoV-19/Denmark/DCGC-7888/2020\|EPI_ISL_618861\|2020 | ···················· | ···················· | ···················· | ···················· | ···················· | ···················· |
| P | 969 | hCoV-19/Denmark/DCGC-7894/2020\|EPI_ISL_618862\|2020 | ···················· | ···················· | ···················· | ···················· | ···················· | ···················· |
| P | 970 | hCoV-19/Denmark/DCGC-7903/2020\|EPI_ISL_618863\|2020 | ···················· | ···················· | ···················· | ···················· | ···················· | ···················· |
| P | 971 | hCoV-19/Denmark/DCGC-7910/2020\|EPI_ISL_618864\|2020 | ···················· | ···················· | ···················· | ···················· | ···················· | ···················· |
| P | 972 | hCoV-19/Denmark/DCGC-7916/2020\|EPI_ISL_618865\|2020 | ···················· | ···················· | ···················· | ···················· | ···················· | ···················· |
| P | 973 | hCoV-19/Denmark/DCGC-7862/2020\|EPI_ISL_618866\|2020 | ···················· | ···················· | ···················· | ···················· | ···················· | ···················· |
| P | 974 | hCoV-19/Denmark/DCGC-7870/2020\|EPI_ISL_618867\|2020 | ···················· | ···················· | ···················· | ···················· | ···················· | ···················· |
| P | 975 | hCoV-19/Denmark/DCGC-7876/2020\|EPI_ISL_618868\|2020 | ···················· | ···················· | ···················· | ···················· | ···················· | ···················· |
| P | 976 | hCoV-19/Denmark/DCGC-7881/2020\|EPI_ISL_618869\|2020 | ···················· | ···················· | ···················· | ···················· | ···················· | ···················· |
| P | 977 | hCoV-19/Denmark/DCGC-7889/2020\|EPI_ISL_618870\|2020 | ···················· | ···················· | ···················· | ···················· | ···················· | ···················· |
| P | 978 | hCoV-19/Denmark/DCGC-7895/2020\|EPI_ISL_618871\|2020 | ···················· | ···················· | ···················· | ···················· | ···················· | ···················· |
| P | 979 | hCoV-19/Denmark/DCGC-7898/2020\|EPI_ISL_618872\|2020 | ···················· | ···················· | ···················· | ···················· | ···················· | ···················· |
| P | 980 | hCoV-19/Denmark/DCGC-7904/2020\|EPI_ISL_618873\|2020 | ···················· | ···················· | ···················· | ···················· | ···················· | ···················· |
| P | 981 | hCoV-19/Denmark/DCGC-7911/2020\|EPI_ISL_618874\|2020 | ···················· | ···················· | ···················· | ···················· | ···················· | ···················· |
| P | 982 | hCoV-19/Denmark/DCGC-7917/2020\|EPI_ISL_618875\|2020 | ···················· | ···················· | ···················· | ···················· | ···················· | ···················· |
| P | 983 | hCoV-19/Denmark/DCGC-7926/2020\|EPI_ISL_618876\|2020 | ···················· | ···················· | ···················· | ···················· | ···················· | ···················· |
| P | 984 | hCoV-19/Denmark/DCGC-7936/2020\|EPI_ISL_618877\|2020 | ···················· | ···················· | ···················· | ···················· | ···················· | ···················· |
| P | 985 | hCoV-19/Denmark/DCGC-7940/2020\|EPI_ISL_618878\|2020 | ···················· | ···················· | ···················· | ···················· | ···················· | ···················· |
| P | 986 | hCoV-19/Denmark/DCGC-7944/2020\|EPI_ISL_618879\|2020 | ···················· | ···················· | ···················· | ···················· | ···················· | ···················· |
| P | 987 | hCoV-19/Denmark/DCGC-7934/2020\|EPI_ISL_618890\|2020 | ···················· | ···················· | ···················· | ···················· | ···················· | ···················· |
| P | 988 | hCoV-19/Denmark/DCGC-7937/2020\|EPI_ISL_618891\|2020 | ···················· | ···················· | ···················· | ···················· | ···················· | ···················· |
| P | 989 | hCoV-19/Denmark/DCGC-7942/2020\|EPI_ISL_618892\|2020 | ···················· | ···················· | ···················· | ···················· | ···················· | ···················· |
| P | 990 | hCoV-19/Denmark/DCGC-7946/2020\|EPI_ISL_618893\|2020 | ···················· | ···················· | ···················· | ···················· | ···················· | ···················· |
| P | 991 | hCoV-19/England/MILK-ABB116/2020\|EPI_ISL_623317\|20 | ···················· | ···················· | ···················· | ···················· | ···················· | ···················· |
| P | 992 | hCoV-19/England/MILK-AB762A/2020\|EPI_ISL_623318\|20 | ···················· | ···················· | ···················· | ···················· | ···················· | ···················· |
| P | 993 | hCoV-19/England/MILK-AB7F6B/2020\|EPI_ISL_623319\|20 | ···················· | ···················· | ···················· | ···················· | ···················· | ···················· |

```
P   994 hCoV-19/England/MILK-AB744E/2020|EPI_ISL_623320|20   ····················  ····················  ·····················  ····················  ····················  ····················
P   995 hCoV-19/England/CAMC-AAFCF6/2020|EPI_ISL_623321|20   ····················  ····················  ·····················  ····················  ······T·············  ····················
P   996 hCoV-19/England/MILK-AB7AB5/2020|EPI_ISL_623322|20   ····················  ····················  ·····················  ····················  ····················  ····················
P   997 hCoV-19/England/ALDP-A95DB4/2020|EPI_ISL_624648|20   ····················  ····················  ·····················  ····················  ····················  ····················
P   998 hCoV-19/England/ALDP-A953B2/2020|EPI_ISL_624651|20   ····················  ····················  ·····················  ····················  ····················  ····················
P   999 hCoV-19/England/ALDP-A945CC/2020|EPI_ISL_624652|20   ····················  ····················  ·····················  ····················  ····················  ····················
P  1000 hCoV-19/Denmark/DCGC-8550/2020|EPI_ISL_625900|2020   ····················  ····················  ·····················  ····················  ····················  ····················
P  1001 hCoV-19/Denmark/DCGC-8504/2020|EPI_ISL_625901|2020   ····················  ····················  ·····················  ····················  ····················  ····················
P  1002 hCoV-19/Denmark/DCGC-8508/2020|EPI_ISL_625902|2020   ····················  ····················  ·····················  ····················  ····················  ····················
P  1003 hCoV-19/Denmark/DCGC-8538/2020|EPI_ISL_625903|2020   ····················  ····················  ·····················  ····················  ····················  ····················
P  1004 hCoV-19/Denmark/DCGC-8551/2020|EPI_ISL_625904|2020   ····················  ····················  ·····················  ····················  ····················  ····················
P  1005 hCoV-19/Denmark/DCGC-8498/2020|EPI_ISL_625905|2020   ····················  ····················  ·····················  ····················  ····················  ····················
P  1006 hCoV-19/Denmark/DCGC-8505/2020|EPI_ISL_625906|2020   ····················  ·A··················  ·····················  ····················  ····················  ····················
P  1007 hCoV-19/Denmark/DCGC-8509/2020|EPI_ISL_625907|2020   ····················  ····················  ·····················  ····················  ····················  ····················
P  1008 hCoV-19/Denmark/DCGC-8516/2020|EPI_ISL_625908|2020   ····················  ····················  ·····················  ····················  ····················  ····················
P  1009 hCoV-19/Denmark/DCGC-8522/2020|EPI_ISL_625909|2020   ····················  ····················  ·····················  ····················  ····················  ····················
P  1010 hCoV-19/Denmark/DCGC-8527/2020|EPI_ISL_625910|2020   ····················  ····················  ·····················  ····················  ····················  ····················
P  1011 hCoV-19/Denmark/DCGC-8534/2020|EPI_ISL_625911|2020   ····················  ····················  ·····················  ····················  ····················  ····················
P  1012 hCoV-19/Denmark/DCGC-8491/2020|EPI_ISL_625912|2020   ····················  ····················  ·····················  ····················  ····················  ····················
P  1013 hCoV-19/Denmark/DCGC-8528/2020|EPI_ISL_625913|2020   ····················  ····················  ·····················  ····················  ····················  ····················
P  1014 hCoV-19/Denmark/DCGC-8539/2020|EPI_ISL_625914|2020   ····················  ····················  ·····················  ····················  ····················  ····················
P  1015 hCoV-19/Denmark/DCGC-8545/2020|EPI_ISL_625915|2020   ····················  ·········A··········  ·····················  ····················  ····················  ····················
P  1016 hCoV-19/Denmark/DCGC-8492/2020|EPI_ISL_625916|2020   ····················  ····················  ·····················  ····················  ····················  ····················
P  1017 hCoV-19/Denmark/DCGC-8499/2020|EPI_ISL_625917|2020   ····················  ····················  ·····················  ····················  ····················  ····················
P  1018 hCoV-19/Denmark/DCGC-8507/2020|EPI_ISL_625918|2020   ····················  ····················  ·····················  ····················  ····················  ····················
P  1019 hCoV-19/Denmark/DCGC-8510/2020|EPI_ISL_625919|2020   ····················  ····················  ·····················  ····················  ····················  ····················
P  1020 hCoV-19/Denmark/DCGC-8513/2020|EPI_ISL_625920|2020   ····················  ····················  ·····················  ····················  ····················  ····················
P  1021 hCoV-19/Denmark/DCGC-8517/2020|EPI_ISL_625921|2020   ····················  ····················  ·····················  ····················  ····················  ····················
P  1022 hCoV-19/Denmark/DCGC-8529/2020|EPI_ISL_625922|2020   ····················  ····················  ·····················  ····················  ····················  ····················
P  1023 hCoV-19/Denmark/DCGC-8540/2020|EPI_ISL_625923|2020   ····················  ····················  ·····················  ····················  ····················  ····················
P  1024 hCoV-19/Denmark/DCGC-8649/2020|EPI_ISL_625982|2020   ····················  ····················  ·····················  ····················  ····················  ····················
P  1025 hCoV-19/Denmark/DCGC-7747/2020|EPI_ISL_618701|2020   ····················  ····················  ·····················  ····················  ····················  ····················
P  1026 hCoV-19/Denmark/DCGC-7762/2020|EPI_ISL_618703|2020   ····················  ····················  ·····················  ····················  ····················  ····················
P  1027 hCoV-19/Denmark/DCGC-7767/2020|EPI_ISL_618704|2020   ····················  ·A··················  ·····················  ····················  ····················  ···A················
P  1028 hCoV-19/Denmark/DCGC-7773/2020|EPI_ISL_618705|2020   ····················  ····················  ·····················  ····················  ····················  ····················
P  1029 hCoV-19/Denmark/DCGC-7778/2020|EPI_ISL_618706|2020   ····················  ····················  ·····················  ····················  ····················  ····················
P  1030 hCoV-19/Denmark/DCGC-7784/2020|EPI_ISL_618707|2020   ····················  ····················  ·····················  ············A·······  ····················  ····················
P  1031 hCoV-19/Denmark/DCGC-7726/2020|EPI_ISL_618709|2020   ····················  ·A··················  ·····················  ····················  ····················  ····················
P  1032 hCoV-19/Denmark/DCGC-7731/2020|EPI_ISL_618710|2020   ····················  ····················  ·····················  ····················  ····················  ····················
P  1033 hCoV-19/Denmark/DCGC-7738/2020|EPI_ISL_618711|2020   ····················  ····················  ·····················  ····················  ····················  ····················
P  1034 hCoV-19/Denmark/DCGC-7748/2020|EPI_ISL_618712|2020   ····················  ····················  ·····················  ····················  ····················  ····················
P  1035 hCoV-19/Denmark/DCGC-7785/2020|EPI_ISL_618714|2020   ····················  ····················  ·····················  ····················  ····················  ····················
P  1036 hCoV-19/Denmark/DCGC-7727/2020|EPI_ISL_618715|2020   ····················  ····················  ·····················  ····················  ····················  ····················
P  1037 hCoV-19/Denmark/DCGC-7739/2020|EPI_ISL_618716|2020   ····················  ····················  ·····················  ····················  ····················  ····················
P  1038 hCoV-19/Denmark/DCGC-7743/2020|EPI_ISL_618717|2020   ····················  ····················  ·····················  ····················  ····················  ····················
P  1039 hCoV-19/Denmark/DCGC-7749/2020|EPI_ISL_618718|2020   ····················  ····················  ·····················  ····················  ····················  ····················
P  1040 hCoV-19/Denmark/DCGC-4180/2020|EPI_ISL_622000|2020   ····················  ····················  ·····················  ····················  ····················  ····················
P  1041 hCoV-19/Denmark/DCGC-4181/2020|EPI_ISL_622001|2020   ····················  ····················  ·····················  ····················  ····················  ····················
P  1042 hCoV-19/Denmark/DCGC-4183/2020|EPI_ISL_622003|2020   ····················  ·A··················  ·····················  ····················  ····················  ···A················
P  1043 hCoV-19/Denmark/DCGC-4184/2020|EPI_ISL_622004|2020   ····················  ····················  ·····················  ····················  ····················  ····················
P  1044 hCoV-19/Denmark/DCGC-4049/2020|EPI_ISL_622005|2020   ····················  ····················  ·····················  ····················  ····················  ····················
P  1045 hCoV-19/Denmark/DCGC-4050/2020|EPI_ISL_622006|2020   ····················  ····················  ·····················  ····················  ····················  ····················
P  1046 hCoV-19/Denmark/DCGC-4051/2020|EPI_ISL_622007|2020   ····················  ····················  ·····················  ····················  ····················  ····················
P  1047 hCoV-19/Denmark/DCGC-4052/2020|EPI_ISL_622008|2020   ····················  ·················A··  ·····················  ····················  ····················  ····················   1
P  1048 hCoV-19/Denmark/DCGC-4054/2020|EPI_ISL_622010|2020   ····················  ····················  ·····················  ····················  ····················  ····················
P  1049 hCoV-19/Denmark/DCGC-4055/2020|EPI_ISL_622011|2020   ····················  ····················  ·····················  ····················  ····················  ····················
P  1050 hCoV-19/Denmark/DCGC-4056/2020|EPI_ISL_622012|2020   ····················  ····················  ·····················  ····················  ····················  ····················
P  1051 hCoV-19/Denmark/DCGC-4058/2020|EPI_ISL_622014|2020   ····················  ····················  ·····················  ····················  ····················  ····················
P  1052 hCoV-19/Denmark/DCGC-4059/2020|EPI_ISL_622015|2020   ····················  ····················  ·····················  ····················  ····················  ····················
P  1053 hCoV-19/Denmark/DCGC-4060/2020|EPI_ISL_622016|2020   ····················  ····················  ·····················  ····················  ····················  ····················
P  1054 hCoV-19/Denmark/DCGC-4062/2020|EPI_ISL_622018|2020   ····················  ····················  ·····················  ····················  ····················  ····················
P  1055 hCoV-19/England/MILK-AB75F0/2020|EPI_ISL_623229|20   ····················  ····················  ·····················  ····················  ····················  ····················
P  1056 hCoV-19/England/MILK-AB8076/2020|EPI_ISL_623230|20   ····················  ····················  ·····················  ····················  ····················  ····················
P  1057 hCoV-19/England/MILK-ABB082/2020|EPI_ISL_623231|20   ····················  ····················  ·····················  ····················  ····················  ····················
P  1058 hCoV-19/England/MILK-ABB91E/2020|EPI_ISL_623232|20   ····················  ····················  ·····················  ····················  ····················  ····················
P  1059 hCoV-19/England/MILK-AB77BE/2020|EPI_ISL_623233|20   ····················  ····················  ·····················  ····················  ····················  ····················
P  1060 hCoV-19/England/MILK-AB8252/2020|EPI_ISL_623236|20   ····················  ····················  ·····················  ····················  ····················  ····················
P  1061 hCoV-19/England/MILK-AB7718/2020|EPI_ISL_623237|20   ····················  ····················  ·····················  ····················  ······T·············  ····················
P  1063 hCoV-19/England/MILK-ABB94B/2020|EPI_ISL_623240|20   ····················  ·A··················  ·····················  ····················  ····················  ···A················
P  1064 hCoV-19/England/MILK-ABB95A/2020|EPI_ISL_623241|20   ····················  ····················  ·····················  ····················  ····················  ····················
P  1065 hCoV-19/England/MILK-ABBAB1/2020|EPI_ISL_623242|20   ····················  ····················  ·····················  ····················  ····················  ····················
P  1066 hCoV-19/England/MILK-ABB3E3/2020|EPI_ISL_623243|20   ····················  ····················  ·····················  ····················  ······T·············  ····················
P  1067 hCoV-19/England/MILK-AB7A88/2020|EPI_ISL_623244|20   ····················  ····················  ·····················  ····················  ····················  ····················
P  1068 hCoV-19/England/MILK-ABB538/2020|EPI_ISL_623246|20   ····················  ····················  ·····················  ····················  ····················  ····················
P  1069 hCoV-19/England/CAMC-AB032A/2020|EPI_ISL_623247|20   ····················  ····················  ·····················  ····················  ····················  ····················
P  1070 hCoV-19/England/CAMC-AAF995/2020|EPI_ISL_623248|20   ····················  ····················  ·····················  ····················  ····················  ····················
P  1071 hCoV-19/England/MILK-AB80A3/2020|EPI_ISL_623249|20   ····················  ····················  ·····················  ····················  ····················  ····················
P  1072 hCoV-19/England/MILK-ABB529/2020|EPI_ISL_623265|20   ····················  ····················  ·····················  ····················  ····················  ····················
P  1073 hCoV-19/England/MILK-ABB7D8/2020|EPI_ISL_623270|20   ····················  ····················  ·····················  ····················  ······T·············  ····················
P  1074 hCoV-19/England/MILK-ABB750/2020|EPI_ISL_623271|20   ····················  ····················  ·····················  ····················  ····················  ····················
```

| | | | | | | | | |
|---|---|---|---|---|---|---|---|---|
| P | 1075 | hCoV-19/England/CAMC-AAF959/2020\|EPI_ISL_623273\|20 | ···················· | ···················· | ····················· | ···················· | ···················· | ···················· |
| P | 1076 | hCoV-19/England/MILK-AB7639/2020\|EPI_ISL_623274\|20 | ···················· | ···················· | ····················· | ···················· | ···················· | ···················· |
| P | 1077 | hCoV-19/England/MILK-AB74D5/2020\|EPI_ISL_623275\|20 | ···················· | ···················· | ····················· | ···················· | ······T············· | ···················· |
| P | 1078 | hCoV-19/England/CAMC-AB0111/2020\|EPI_ISL_623276\|20 | ···················· | ···················· | ····················· | ···················· | ···················· | ···················· |
| P | 1079 | hCoV-19/England/MILK-AB7AA6/2020\|EPI_ISL_623277\|20 | ···················· | ···················· | ····················· | ···················· | ···················· | ···················· |
| P | 1080 | hCoV-19/England/MILK-AB75E1/2020\|EPI_ISL_623279\|20 | ···················· | ···················· | ····················· | ···················· | ···················· | ···················· |
| P | 1081 | hCoV-19/England/MILK-AB760C/2020\|EPI_ISL_623280\|20 | ···················· | ···················· | ····················· | ···················· | ···················· | ···················· |
| P | 1082 | hCoV-19/England/CAMC-AB0393/2020\|EPI_ISL_623281\|20 | ···················· | ···················· | ····················· | ···················· | ···················· | ···················· |
| P | 1083 | hCoV-19/England/MILK-AB82DA/2020\|EPI_ISL_623282\|20 | ···················· | ···················· | ····················· | ···················· | ···················· | ···················· |
| P | 1084 | hCoV-19/England/MILK-AB7648/2020\|EPI_ISL_623283\|20 | ···················· | ···················· | ····················· | ···················· | ···················· | ···················· |
| P | 1085 | hCoV-19/England/MILK-ABB617/2020\|EPI_ISL_623284\|20 | ···················· | ···················· | ····················· | ···················· | ···················· | ···················· |
| P | 1086 | hCoV-19/England/MILK-AB802B/2020\|EPI_ISL_623285\|20 | ···················· | ···················· | ····················· | ···················· | ···················· | ···················· |
| P | 1087 | hCoV-19/England/CAMC-AB014E/2020\|EPI_ISL_623286\|20 | ···················· | ···················· | ····················· | ···················· | ···················· | ···················· |
| P | 1088 | hCoV-19/England/MILK-AB800D/2020\|EPI_ISL_623287\|20 | ···················· | ···················· | ····················· | ···················· | ······T············· | ···················· |
| P | 1089 | hCoV-19/England/CAMC-AAFD11/2020\|EPI_ISL_623288\|20 | ···················· | ···················· | ····················· | ···················· | ······T············· | ···················· |
| P | 1090 | hCoV-19/England/MILK-ABB41D/2020\|EPI_ISL_623289\|20 | ···················· | ···················· | ····················· | ···················· | ···················· | ···················· |
| P | 1091 | hCoV-19/England/MILK-AB73E7/2020\|EPI_ISL_623290\|20 | ···················· | ···················· | ····················· | ···················· | ···················· | ···················· |
| P | 1092 | hCoV-19/England/MILK-AB77FA/2020\|EPI_ISL_623291\|20 | ···················· | ···················· | ····················· | ···················· | ···················· | ···················· |
| P | 1093 | hCoV-19/England/MILK-AB73F6/2020\|EPI_ISL_623292\|20 | ···················· | ···················· | ····················· | ···················· | ···················· | ···················· |
| P | 1094 | hCoV-19/England/MILK-AB7763/2020\|EPI_ISL_623293\|20 | ···················· | ···················· | ····················· | ···················· | ···················· | ···················· |
| P | 1095 | hCoV-19/England/MILK-ABB2AA/2020\|EPI_ISL_623294\|20 | ···················· | ···················· | ····················· | ···················· | ···················· | ···················· |
| P | 1096 | hCoV-19/England/MILK-ABB0CE/2020\|EPI_ISL_623296\|20 | ···················· | ···················· | ····················· | ···················· | ···················· | ···················· |
| P | 1097 | hCoV-19/England/MILK-AB7A2E/2020\|EPI_ISL_623297\|20 | ···················· | ···················· | ····················· | ···················· | ···················· | ···················· |
| P | 1098 | hCoV-19/England/MILK-AB747B/2020\|EPI_ISL_623299\|20 | ···················· | ···················· | ····················· | ···················· | ···················· | ···················· |
| P | 1099 | hCoV-19/England/MILK-AB8085/2020\|EPI_ISL_623300\|20 | ···················· | ···················· | ····················· | ···················· | ···················· | ···················· |
| P | 1100 | hCoV-19/England/CAMC-AB0339/2020\|EPI_ISL_623301\|20 | ···················· | ···················· | ····················· | ···················· | ···················· | ···················· |
| P | 1101 | hCoV-19/England/MILK-ABB42C/2020\|EPI_ISL_623302\|20 | ···················· | ···················· | ····················· | ···················· | ···················· | ···················· |
| P | 1102 | hCoV-19/England/MILK-AB83F5/2020\|EPI_ISL_623303\|20 | ···················· | ···················· | ····················· | ···················· | ···················· | ···················· |
| P | 1103 | hCoV-19/England/MILK-ABB0EC/2020\|EPI_ISL_623304\|20 | ···················· | ···················· | ····················· | ···················· | ···················· | ···················· |
| P | 1104 | hCoV-19/England/MILK-ABB8C6/2020\|EPI_ISL_623305\|20 | ···················· | ···················· | ····················· | ···················· | ···················· | ···················· |
| P | 1105 | hCoV-19/England/MILK-AB838C/2020\|EPI_ISL_623306\|20 | ···················· | ·A·················· | ····················· | ···················· | ···················· | ···A················ |
| P | 1106 | hCoV-19/England/MILK-AB82F8/2020\|EPI_ISL_623307\|20 | ···················· | ···················· | ····················· | ···················· | ···················· | ···················· |
| P | 1107 | hCoV-19/England/CAMC-AAFCD8/2020\|EPI_ISL_623308\|20 | ···················· | ···················· | ····················· | ···················· | ···················· | ···················· |
| P | 1108 | hCoV-19/England/MILK-AB7666/2020\|EPI_ISL_623309\|20 | ···················· | ···················· | ····················· | ···················· | ···················· | ···················· |
| P | 1109 | hCoV-19/England/MILK-ABB6AE/2020\|EPI_ISL_623310\|20 | ···················· | ···················· | ····················· | ···················· | ···················· | ···················· |
| P | 1110 | hCoV-19/England/CAMC-AAFEB4/2020\|EPI_ISL_623312\|20 | ···················· | ···················· | ····················· | ···················· | ···················· | ···················· |
| P | 1111 | hCoV-19/England/MILK-AB7657/2020\|EPI_ISL_623314\|20 | ···················· | ···················· | ····················· | ···················· | ···················· | ···················· |
| P | 1112 | hCoV-19/England/MILK-AB7693/2020\|EPI_ISL_623315\|20 | ···················· | ···················· | ····················· | ···················· | ···················· | ···················· |
| P | 1113 | hCoV-19/England/MILK-AB7E22/2020\|EPI_ISL_623316\|20 | ···················· | ···················· | ····················· | ···················· | ···················· | ···················· |
| P | 1114 | hCoV-19/England/CAMC-AAFDC6/2020\|EPI_ISL_623323\|20 | ···················· | ···················· | ····················· | ···················· | ···················· | ···················· |
| P | 1115 | hCoV-19/England/MILK-AB8067/2020\|EPI_ISL_623324\|20 | ···················· | ···················· | ····················· | ···················· | ···················· | ···················· |
| P | 1116 | hCoV-19/England/MILK-AB7E6E/2020\|EPI_ISL_623327\|20 | ···················· | ···················· | ····················· | ···················· | ···················· | ···················· |
| P | 1117 | hCoV-19/England/MILK-ABB626/2020\|EPI_ISL_623328\|20 | ···················· | ···················· | ····················· | ···················· | ···················· | ···················· |
| P | 1118 | hCoV-19/England/MILK-AB86DE/2020\|EPI_ISL_623329\|20 | ···················· | ···················· | ····················· | ···················· | ···················· | ···················· |
| P | 1119 | hCoV-19/England/CAMC-AB02E1/2020\|EPI_ISL_623330\|20 | ···················· | ···················· | ····················· | ···················· | ···················· | ···················· |
| P | 1120 | hCoV-19/England/MILK-AB7736/2020\|EPI_ISL_623331\|20 | ···················· | ···················· | ····················· | ···················· | ···················· | ···················· |
| P | 1121 | hCoV-19/England/MILK-AB839B/2020\|EPI_ISL_623332\|20 | ···················· | ···················· | ····················· | ···················· | ···················· | ···················· |
| P | 1122 | hCoV-19/England/CAMC-AAFC32/2020\|EPI_ISL_623333\|20 | ···················· | ···················· | ····················· | ···················· | ···················· | ···················· |
| P | 1123 | hCoV-19/England/CAMC-AB013F/2020\|EPI_ISL_623334\|20 | ···················· | ···················· | ····················· | ···················· | ···················· | ···················· |
| P | 1124 | hCoV-19/England/MILK-ABB7AB/2020\|EPI_ISL_623335\|20 | ···················· | ···················· | ····················· | ···················· | ···················· | ···················· |
| P | 1125 | hCoV-19/England/MILK-AB8304/2020\|EPI_ISL_623336\|20 | ···················· | ···················· | ····················· | ···················· | ···················· | ···················· |
| P | 1126 | hCoV-19/England/MILK-AB7727/2020\|EPI_ISL_623338\|20 | ···················· | ···················· | ····················· | ···················· | ···················· | ···················· |
| P | 1127 | hCoV-19/England/MILK-AB73AB/2020\|EPI_ISL_623339\|20 | ···················· | ···················· | ····················· | ···················· | ···················· | ···················· |
| P | 1128 | hCoV-19/England/MILK-AB76B1/2020\|EPI_ISL_623340\|20 | ···················· | ···················· | ····················· | ···················· | ···················· | ···················· |
| P | 1129 | hCoV-19/England/MILK-AB7DDA/2020\|EPI_ISL_623341\|20 | ···················· | ···················· | ····················· | ···················· | ···················· | ···················· |
| P | 1130 | hCoV-19/England/MILK-AB83B9/2020\|EPI_ISL_623342\|20 | ···················· | ···················· | ····················· | ···················· | ···················· | ···················· |
| P | 1131 | hCoV-19/England/CAMC-AAFE87/2020\|EPI_ISL_623344\|20 | ···················· | ···················· | ····················· | ···················· | ······T············· | ···················· |
| P | 1132 | hCoV-19/England/CAMC-AAFC41/2020\|EPI_ISL_623345\|20 | ···················· | ···················· | ····················· | ···················· | ···················· | ···················· |
| P | 1133 | hCoV-19/England/CAMC-AAFDB7/2020\|EPI_ISL_623346\|20 | ···················· | ···················· | ····················· | ···················· | ···················· | ···················· |
| P | 1134 | hCoV-19/England/CAMC-AB0384/2020\|EPI_ISL_623347\|20 | ···················· | ···················· | ····················· | ···················· | ···················· | ···················· |
| P | 1135 | hCoV-19/England/ALDP-A959FC/2020\|EPI_ISL_624560\|20 | ···················· | ···················· | ····················· | ···················· | ···················· | ···················· |
| P | 1136 | hCoV-19/England/ALDP-A95E66/2020\|EPI_ISL_624561\|20 | ···················· | ···················· | ····················· | ···················· | ···················· | ···················· |
| P | 1137 | hCoV-19/England/ALDP-A95A08/2020\|EPI_ISL_624563\|20 | ···················· | ···················· | ····················· | ···················· | ···················· | ···················· |
| P | 1138 | hCoV-19/England/ALDP-A96454/2020\|EPI_ISL_624564\|20 | ···················· | ···················· | ····················· | ···················· | ···················· | ···················· |
| P | 1139 | hCoV-19/England/ALDP-A94F55/2020\|EPI_ISL_624565\|20 | ···················· | ···················· | ····················· | ···················· | ···················· | ···················· |
| P | 1140 | hCoV-19/England/ALDP-A95DA5/2020\|EPI_ISL_624566\|20 | ···················· | ···················· | ····················· | ···················· | ···················· | ···················· |
| P | 1141 | hCoV-19/England/ALDP-A94CE5/2020\|EPI_ISL_624567\|20 | ···················· | ···················· | ····················· | ···················· | ···················· | ···················· |
| P | 1142 | hCoV-19/England/ALDP-A94C21/2020\|EPI_ISL_624568\|20 | ···················· | ···················· | ····················· | ···················· | ···················· | ···················· |
| P | 1143 | hCoV-19/England/ALDP-A945BD/2020\|EPI_ISL_624569\|20 | ···················· | ···················· | ····················· | ···················· | ···················· | ···················· |
| P | 1144 | hCoV-19/England/ALDP-A964BE/2020\|EPI_ISL_624570\|20 | ···················· | ···················· | ····················· | ···················· | ···················· | ···················· |
| P | 1145 | hCoV-19/England/ALDP-A94298/2020\|EPI_ISL_624571\|20 | ···················· | ···················· | ····················· | ···················· | ···················· | ···················· |
| P | 1146 | hCoV-19/England/ALDP-A940CB/2020\|EPI_ISL_624572\|20 | ···················· | ···················· | ····················· | ···················· | ···················· | ···················· |
| P | 1147 | hCoV-19/England/ALDP-A9530D/2020\|EPI_ISL_624573\|20 | ···················· | ···················· | ····················· | ···················· | ···················· | ···················· |
| P | 1148 | hCoV-19/England/ALDP-A945AE/2020\|EPI_ISL_624574\|20 | ···················· | ···················· | ····················· | ···················· | ···················· | ···················· |
| P | 1149 | hCoV-19/England/ALDP-A95D78/2020\|EPI_ISL_624575\|20 | ···················· | ···················· | ····················· | ···················· | ······T············· | ···············A···· |
| P | 1150 | hCoV-19/England/ALDP-A9473F/2020\|EPI_ISL_624576\|20 | ···················· | ···················· | ····················· | ···················· | ···················· | ···················· |
| P | 1151 | hCoV-19/England/ALDP-A94562/2020\|EPI_ISL_624578\|20 | ···················· | ·A·················· | ····················· | ···················· | ···················· | ···················· |
| P | 1152 | hCoV-19/England/ALDP-A95279/2020\|EPI_ISL_624579\|20 | ···················· | ···················· | ····················· | ···················· | ···················· | ···················· |
| P | 1153 | hCoV-19/England/ALDP-A9558F/2020\|EPI_ISL_624596\|20 | ···················· | ···················· | ····················· | ···················· | ···················· | ···················· |
| P | 1154 | hCoV-19/England/ALDP-A96348/2020\|EPI_ISL_624597\|20 | ···················· | ···················· | ····················· | ···················· | ···················· | ···················· |

| | | | | | | | | |
|---|---|---|---|---|---|---|---|---|
| P | 1155 | hCoV-19/England/ALDP-A96023/2020\|EPI_ISL_624600\|20 | ···················· | ···················· | ···················· | ···················· | ···················· | ···················· |
| P | 1156 | hCoV-19/England/ALDP-A96296/2020\|EPI_ISL_624601\|20 | ···················· | ···················· | ···················· | ···················· | ···················· | ···················· |
| P | 1157 | hCoV-19/England/ALDP-A945DB/2020\|EPI_ISL_624603\|20 | ···················· | ···················· | ···················· | ···················· | ···················· | ···················· |
| P | 1158 | hCoV-19/England/ALDP-A94711/2020\|EPI_ISL_624604\|20 | ···················· | ···················· | ···················· | ···················· | ···················· | ···················· |
| P | 1159 | hCoV-19/England/ALDP-A95A17/2020\|EPI_ISL_624605\|20 | ···················· | ···················· | ···················· | ···················· | ···················· | ···················· |
| P | 1160 | hCoV-19/England/ALDP-A94CB8/2020\|EPI_ISL_624606\|20 | ···················· | ···················· | ···················· | ···················· | ···················· | ···················· |
| P | 1161 | hCoV-19/England/ALDP-A952D3/2020\|EPI_ISL_624607\|20 | ···················· | ···················· | ···················· | ···················· | ···················· | ···················· |
| P | 1162 | hCoV-19/England/ALDP-A95FCD/2020\|EPI_ISL_624608\|20 | ···················· | ···················· | ···················· | ···················· | ···················· | ···················· |
| P | 1163 | hCoV-19/England/ALDP-A963B1/2020\|EPI_ISL_624609\|20 | ···················· | ···················· | ···················· | ···················· | ···················· | ···················· |
| P | 1164 | hCoV-19/England/ALDP-A96366/2020\|EPI_ISL_624610\|20 | ···················· | ···················· | ···················· | ···················· | ···················· | ···················· |
| P | 1165 | hCoV-19/England/ALDP-A94F82/2020\|EPI_ISL_624611\|20 | ···················· | ···················· | ···················· | ···················· | ···················· | ···················· |
| P | 1166 | hCoV-19/England/ALDP-A9606F/2020\|EPI_ISL_624612\|20 | ···················· | ···················· | ···················· | ···················· | ···················· | ···················· |
| P | 1167 | hCoV-19/England/ALDP-A9425C/2020\|EPI_ISL_624614\|20 | ···················· | ···················· | ···················· | ···················· | ···················· | ···················· |
| P | 1168 | hCoV-19/England/ALDP-A962B4/2020\|EPI_ISL_624615\|20 | ···················· | ···················· | ···················· | ···················· | ···················· | ···················· |
| P | 1169 | hCoV-19/England/ALDP-A96481/2020\|EPI_ISL_624616\|20 | ···················· | ···················· | ···················· | ···················· | ···················· | ···················· |
| P | 1170 | hCoV-19/England/ALDP-A944ED/2020\|EPI_ISL_624620\|20 | ···················· | ···················· | ···················· | ···················· | ···················· | ···················· |
| P | 1171 | hCoV-19/England/ALDP-A95DE1/2020\|EPI_ISL_624621\|20 | ···················· | ···················· | ···················· | ···················· | ···················· | ···················· |
| P | 1172 | hCoV-19/England/ALDP-A9521F/2020\|EPI_ISL_624622\|20 | ···················· | ···················· | ···················· | ···················· | ···················· | ···················· |
| P | 1173 | hCoV-19/England/ALDP-A96490/2020\|EPI_ISL_624623\|20 | ···················· | ···················· | ···················· | ···················· | ···················· | ···················· |
| P | 1174 | hCoV-19/England/ALDP-A958E0/2020\|EPI_ISL_624624\|20 | ···················· | ···················· | ···················· | ···················· | ···················· | ···················· |
| P | 1175 | hCoV-19/England/ALDP-A9623C/2020\|EPI_ISL_624625\|20 | ···················· | ···················· | ···················· | ···················· | ···················· | ···················· |
| P | 1176 | hCoV-19/England/ALDP-A945EA/2020\|EPI_ISL_624626\|20 | ···················· | ···················· | ···················· | ···················· | ···················· | ···················· |
| P | 1177 | hCoV-19/England/ALDP-A94ADC/2020\|EPI_ISL_624627\|20 | ···················· | ···················· | ···················· | ···················· | ···················· | ···················· |
| P | 1178 | hCoV-19/England/ALDP-A96199/2020\|EPI_ISL_624628\|20 | ···················· | ···················· | ···················· | ···················· | ···················· | ···················· |
| P | 1179 | hCoV-19/England/ALDP-A9621E/2020\|EPI_ISL_624630\|20 | ···················· | ···················· | ···················· | ···················· | ···················· | ···················· |
| P | 1180 | hCoV-19/England/ALDP-A963C0/2020\|EPI_ISL_624631\|20 | ···················· | ···················· | ···················· | ···················· | ···················· | ···················· |
| P | 1181 | hCoV-19/England/ALDP-A96506/2020\|EPI_ISL_624632\|20 | ···················· | ···················· | ···················· | ···················· | ···················· | ···················· |
| P | 1182 | hCoV-19/England/ALDP-A963DF/2020\|EPI_ISL_624633\|20 | ···················· | ···················· | ···················· | ···················· | ···················· | ···················· |
| P | 1183 | hCoV-19/England/ALDP-A94ABE/2020\|EPI_ISL_624635\|20 | ···················· | ···················· | ···················· | ···················· | ···················· | ···················· |
| P | 1184 | hCoV-19/England/ALDP-A95A80/2020\|EPI_ISL_624637\|20 | ···················· | ···················· | ···················· | ···················· | ···················· | ···················· |
| P | 1185 | hCoV-19/England/ALDP-A96418/2020\|EPI_ISL_624638\|20 | ···················· | ···················· | ···················· | ···················· | ···················· | ···················· |
| P | 1186 | hCoV-19/England/ALDP-A952F1/2020\|EPI_ISL_624639\|20 | ···················· | ···················· | ···················· | ···················· | ···················· | ···················· |
| P | 1187 | hCoV-19/England/ALDP-A955DA/2020\|EPI_ISL_624640\|20 | ···················· | ···················· | ···················· | ···················· | ·····T·············· | ···················· |
| P | 1188 | hCoV-19/England/ALDP-A953C1/2020\|EPI_ISL_624641\|20 | ···················· | ···················· | ···················· | ···················· | ···················· | ···················· |
| P | 1189 | hCoV-19/England/ALDP-A94456/2020\|EPI_ISL_624642\|20 | ···················· | ···················· | ···················· | ···················· | ···················· | ···················· |
| P | 1190 | hCoV-19/England/ALDP-A95A26/2020\|EPI_ISL_624643\|20 | ···················· | ···················· | ···················· | ···················· | ···················· | ···················· |
| P | 1191 | hCoV-19/England/ALDP-A95FDC/2020\|EPI_ISL_624644\|20 | ···················· | ···················· | ···················· | ···················· | ···················· | ···················· |
| P | 1192 | hCoV-19/England/ALDP-A94EC1/2020\|EPI_ISL_624647\|20 | ···················· | ···················· | ···················· | ···················· | ···················· | ···················· |
| P | 1193 | hCoV-19/England/ALDP-A947D5/2020\|EPI_ISL_624650\|20 | ···················· | ···················· | ···················· | ···················· | ···················· | ···················· |
| P | 1194 | hCoV-19/England/ALDP-A94474/2020\|EPI_ISL_624653\|20 | ···················· | ···················· | ···················· | ···················· | ···················· | ···················· |
| P | 1195 | hCoV-19/England/ALDP-A95F36/2020\|EPI_ISL_624654\|20 | ···················· | ···················· | ···················· | ···················· | ···················· | ···················· |
| P | 1196 | hCoV-19/England/ALDP-A96050/2020\|EPI_ISL_624655\|20 | ···················· | ···················· | ···················· | ···················· | ···················· | ···················· |
| P | 1197 | hCoV-19/England/ALDP-A95E93/2020\|EPI_ISL_624656\|20 | ···················· | ···················· | ···················· | ···················· | ···················· | ···················· |
| P | 1198 | hCoV-19/England/ALDP-A96111/2020\|EPI_ISL_624658\|20 | ···················· | ···················· | ···················· | ···················· | ···················· | ···················· |
| P | 1199 | hCoV-19/England/ALDP-A95A62/2020\|EPI_ISL_624659\|20 | ···················· | ···················· | ···················· | ···················· | ···················· | ···················· |
| P | 1200 | hCoV-19/England/ALDP-A955E9/2020\|EPI_ISL_624660\|20 | ···················· | ···················· | ···················· | ···················· | ···················· | ···················· |
| P | 1201 | hCoV-19/England/ALDP-A95E0C/2020\|EPI_ISL_624661\|20 | ···················· | ···················· | ···················· | ···················· | ···················· | ···················· |
| P | 1202 | hCoV-19/England/ALDP-A94A27/2020\|EPI_ISL_624662\|20 | ···················· | ···················· | ···················· | ···················· | ···················· | ···················· |
| P | 1203 | hCoV-19/England/ALDP-A95AAE/2020\|EPI_ISL_624663\|20 | ···················· | ···················· | ···················· | ···················· | ···················· | ···················· |
| P | 1204 | hCoV-19/England/ALDP-A94517/2020\|EPI_ISL_624664\|20 | ···················· | ···················· | ···················· | ···················· | ···················· | ···················· |
| P | 1205 | hCoV-19/England/ALDP-A94113/2020\|EPI_ISL_624665\|20 | ···················· | ···················· | ···················· | ···················· | ···················· | ···················· |
| P | 1206 | hCoV-19/England/ALDP-A952B5/2020\|EPI_ISL_624666\|20 | ···················· | ···················· | ···················· | ···················· | ···················· | ···················· |
| P | 1207 | hCoV-19/England/ALDP-A95F54/2020\|EPI_ISL_624667\|20 | ···················· | ···················· | ···················· | ···················· | ···················· | ···················· |
| P | 1208 | hCoV-19/England/ALDP-A963A2/2020\|EPI_ISL_624668\|20 | ···················· | ···················· | ···················· | ···················· | ···················· | ···················· |
| P | 1209 | hCoV-19/England/ALDP-A95DD2/2020\|EPI_ISL_624669\|20 | ···················· | ···················· | ···················· | ···················· | ···················· | ···················· |
| P | 1210 | hCoV-19/England/ALDP-A963EE/2020\|EPI_ISL_624670\|20 | ···················· | ···················· | ···················· | ···················· | ···················· | ···················· |
| P | 1211 | hCoV-19/England/ALDP-A94EEF/2020\|EPI_ISL_624672\|20 | ···················· | ···················· | ···················· | ···················· | ···················· | ···················· |
| P | 1212 | hCoV-19/England/ALDP-A941AA/2020\|EPI_ISL_624673\|20 | ···················· | ···················· | ···················· | ···················· | ···················· | ···················· |
| P | 1213 | hCoV-19/England/ALDP-A94EFE/2020\|EPI_ISL_624674\|20 | ···················· | ···················· | ···················· | ···················· | ···················· | ···················· |
| P | 1214 | hCoV-19/England/ALDP-A9477B/2020\|EPI_ISL_624675\|20 | ···················· | ···················· | ···················· | ···················· | ···················· | ···················· |
| P | 1215 | hCoV-19/England/ALDP-A94B7F/2020\|EPI_ISL_624676\|20 | ···················· | ···················· | ···················· | ···················· | ···················· | ···················· |
| P | 1216 | hCoV-19/England/ALDP-A957D4/2020\|EPI_ISL_624677\|20 | ···················· | ···················· | ···················· | ···················· | ···················· | ···················· |
| P | 1217 | hCoV-19/England/ALDP-A961D5/2020\|EPI_ISL_624678\|20 | ···················· | ···················· | ···················· | ···················· | ···················· | ···················· |
| P | 1218 | hCoV-19/England/ALDP-A955BC/2020\|EPI_ISL_624679\|20 | ···················· | ···················· | ···················· | ···················· | ···················· | ···················· |
| P | 1219 | hCoV-19/Denmark/DCGC-8489/2020\|EPI_ISL_625892\|2020 | ···················· | ·A·················· | ···················· | ···················· | ···················· | ···················· |
| P | 1220 | hCoV-19/Denmark/DCGC-8501/2020\|EPI_ISL_625893\|2020 | ···················· | ·A·················· | ···················· | ···················· | ···················· | ···················· |
| P | 1221 | hCoV-19/Denmark/DCGC-8541/2020\|EPI_ISL_625894\|2020 | ···················· | ···················· | ···················· | ···················· | ···················· | ···················· |
| P | 1222 | hCoV-19/Denmark/DCGC-8520/2020\|EPI_ISL_625895\|2020 | ···················· | ···················· | ···················· | ···················· | ···················· | ···················· |
| P | 1223 | hCoV-19/Denmark/DCGC-8536/2020\|EPI_ISL_625896\|2020 | ···················· | ···················· | ···················· | ···················· | ···················· | ···················· |
| P | 1224 | hCoV-19/Denmark/DCGC-8549/2020\|EPI_ISL_625897\|2020 | ···················· | ···················· | ···················· | ···················· | ···················· | ···················· |
| P | 1225 | hCoV-19/Denmark/DCGC-8503/2020\|EPI_ISL_625898\|2020 | ···················· | ···················· | ···················· | ···················· | ···················· | ···················· |
| P | 1226 | hCoV-19/Denmark/DCGC-8543/2020\|EPI_ISL_625899\|2020 | ···················· | ···················· | ···················· | ···················· | ···················· | ···················· |
| P | 1227 | hCoV-19/Denmark/DCGC-8493/2020\|EPI_ISL_625924\|2020 | ···················· | ···················· | ···················· | ···················· | ·····T·············· | ···················· |
| P | 1228 | hCoV-19/Denmark/DCGC-8500/2020\|EPI_ISL_625925\|2020 | ···················· | ···················· | ···················· | ···················· | ···················· | ···················· |
| P | 1229 | hCoV-19/Denmark/DCGC-8514/2020\|EPI_ISL_625926\|2020 | ···················· | ···················· | ···················· | ···················· | ···················· | ···················· |
| P | 1230 | hCoV-19/Denmark/DCGC-8518/2020\|EPI_ISL_625927\|2020 | ···················· | ···················· | ···················· | ···················· | ···················· | ···················· |
| P | 1231 | hCoV-19/Denmark/DCGC-8523/2020\|EPI_ISL_625928\|2020 | ···················· | ···················· | ···················· | ···················· | ···················· | ···················· |
| P | 1232 | hCoV-19/Denmark/DCGC-8530/2020\|EPI_ISL_625929\|2020 | ···················· | ···················· | ···················· | ···················· | ···················· | ···················· |
| P | 1233 | hCoV-19/Denmark/DCGC-8557/2020\|EPI_ISL_625930\|2020 | ···················· | ···················· | ···················· | ···················· | ···················· | ···················· |
| P | 1234 | hCoV-19/Denmark/DCGC-8563/2020\|EPI_ISL_625931\|2020 | ···················· | ···················· | ···················· | ···················· | ···················· | ···················· |

| | | | | | | | |
|---|---|---|---|---|---|---|---|
| P | 1235 hCoV-19/Denmark/DCGC-8576/2020\|EPI_ISL_625932\|2020 | ···················· | ···················· | ···················· | ···················· | ···················· | ···················· |
| P | 1236 hCoV-19/Denmark/DCGC-8602/2020\|EPI_ISL_625934\|2020 | ···················· | ·A·················· | ···················· | ···················· | ···················· | ···A················ |
| P | 1237 hCoV-19/Denmark/DCGC-8553/2020\|EPI_ISL_625935\|2020 | ···················· | ···················· | ···················· | ···················· | ···················· | ···················· |
| P | 1238 hCoV-19/Denmark/DCGC-8577/2020\|EPI_ISL_625936\|2020 | ···················· | ···················· | ···················· | ···················· | ······T············· | ···················· |
| P | 1239 hCoV-19/Denmark/DCGC-8560/2020\|EPI_ISL_625939\|2020 | ···················· | ···················· | ···················· | ···················· | ···················· | ···················· |
| P | 1240 hCoV-19/Denmark/DCGC-8565/2020\|EPI_ISL_625940\|2020 | ···················· | ···················· | ···················· | ···················· | ···················· | ···················· |
| P | 1241 hCoV-19/Denmark/DCGC-8571/2020\|EPI_ISL_625941\|2020 | ···················· | ···················· | ···················· | ···················· | ···················· | ···················· |
| P | 1242 hCoV-19/Denmark/DCGC-8578/2020\|EPI_ISL_625942\|2020 | ···················· | ···················· | ···················· | ···················· | ···················· | ···················· |
| P | 1243 hCoV-19/Denmark/DCGC-8593/2020\|EPI_ISL_625945\|2020 | ···················· | ···················· | ···················· | ···················· | ···················· | ···················· |
| P | 1244 hCoV-19/Denmark/DCGC-8598/2020\|EPI_ISL_625946\|2020 | ···················· | ···················· | ···················· | ···················· | ···················· | ···················· |
| P | 1245 hCoV-19/Denmark/DCGC-8555/2020\|EPI_ISL_625947\|2020 | ···················· | ···················· | ···················· | ···················· | ···················· | ···················· |
| P | 1246 hCoV-19/Denmark/DCGC-8561/2020\|EPI_ISL_625948\|2020 | ···················· | ···················· | ···················· | ···················· | ···················· | ···················· |
| P | 1247 hCoV-19/Denmark/DCGC-8566/2020\|EPI_ISL_625949\|2020 | ···················· | ···················· | ···················· | ···················· | ···················· | ···················· |
| P | 1248 hCoV-19/Denmark/DCGC-8572/2020\|EPI_ISL_625950\|2020 | ···················· | ···················· | ···················· | ···················· | ···················· | ···················· |
| P | 1249 hCoV-19/Denmark/DCGC-8584/2020\|EPI_ISL_625951\|2020 | ········T··········· | ···················· | ···················· | ···················· | ···················· | ···················· |
| P | 1250 hCoV-19/Denmark/DCGC-8567/2020\|EPI_ISL_625953\|2020 | ···················· | ···················· | ···················· | ···················· | ···················· | ···················· |
| P | 1251 hCoV-19/Denmark/DCGC-8573/2020\|EPI_ISL_625954\|2020 | ···················· | ···················· | ···················· | ···················· | ···················· | ···················· |
| P | 1252 hCoV-19/Denmark/DCGC-8562/2020\|EPI_ISL_625956\|2020 | ···················· | ·A·················· | ···················· | ···················· | ···················· | ···················· |
| P | 1253 hCoV-19/Denmark/DCGC-8574/2020\|EPI_ISL_625957\|2020 | ···················· | ···················· | ···················· | ···················· | ···················· | ···················· |
| P | 1254 hCoV-19/Denmark/DCGC-8595/2020\|EPI_ISL_625958\|2020 | ···················· | ···················· | ···················· | ···················· | ···················· | ···················· |
| P | 1255 hCoV-19/Denmark/DCGC-8601/2020\|EPI_ISL_625959\|2020 | ···················· | ···················· | ···················· | ···················· | ······T············· | ···················· |
| P | 1256 hCoV-19/Denmark/DCGC-8569/2020\|EPI_ISL_625961\|2020 | ···················· | ···················· | ···················· | ···················· | ···················· | ···················· |
| P | 1257 hCoV-19/Denmark/DCGC-8575/2020\|EPI_ISL_625962\|2020 | ···················· | ···················· | ···················· | ···················· | ···················· | ···················· |
| P | 1258 hCoV-19/Denmark/DCGC-8587/2020\|EPI_ISL_625963\|2020 | ···················· | ···················· | ···················· | ···················· | ···················· | ···················· |
| P | 1259 hCoV-19/Denmark/DCGC-8596/2020\|EPI_ISL_625964\|2020 | ···················· | ···················· | ···················· | ···················· | ···················· | ···················· |
| P | 1260 hCoV-19/Denmark/DCGC-8606/2020\|EPI_ISL_625965\|2020 | ···················· | ···················· | ···················· | ···················· | ···················· | ···················· |
| P | 1261 hCoV-19/Denmark/DCGC-8619/2020\|EPI_ISL_625966\|2020 | ···················· | ···················· | ···················· | ···················· | ···················· | ···················· |
| P | 1262 hCoV-19/Denmark/DCGC-8666/2020\|EPI_ISL_625969\|2020 | ···················· | ·A·················· | ···················· | ···················· | ···················· | ···················· |
| P | 1263 hCoV-19/Denmark/DCGC-8607/2020\|EPI_ISL_625971\|2020 | ···················· | ···················· | ···················· | ···················· | ···················· | ···················· |
| P | 1264 hCoV-19/Denmark/DCGC-8662/2020\|EPI_ISL_625976\|2020 | ···················· | ···················· | ···················· | ···················· | ···················· | ···················· |
| P | 1265 hCoV-19/Denmark/DCGC-8667/2020\|EPI_ISL_625977\|2020 | ···················· | ···················· | ···················· | ···················· | ···················· | ···················· |
| P | 1266 hCoV-19/Denmark/DCGC-8608/2020\|EPI_ISL_625978\|2020 | ···················· | ···················· | ···················· | ···················· | ···················· | ···················· |
| P | 1267 hCoV-19/Denmark/DCGC-8638/2020\|EPI_ISL_625981\|2020 | ···················· | ·A·················· | ···················· | ···················· | ···················· | ···················· |
| P | 1268 hCoV-19/Denmark/DCGC-8615/2020\|EPI_ISL_625985\|2020 | ···················· | ···················· | ···················· | ···················· | ···················· | ···················· |
| P | 1269 hCoV-19/Denmark/DCGC-8622/2020\|EPI_ISL_625986\|2020 | ···················· | ···················· | ···················· | ···················· | ···················· | ···················· |
| P | 1270 hCoV-19/Denmark/DCGC-8639/2020\|EPI_ISL_625987\|2020 | ···················· | ···················· | ···················· | ···················· | ···················· | ···················· |
| P | 1271 hCoV-19/Denmark/DCGC-8669/2020\|EPI_ISL_625988\|2020 | ···················· | ···················· | ···················· | ···················· | ···················· | ···················· |
| P | 1272 hCoV-19/Denmark/DCGC-8610/2020\|EPI_ISL_625990\|2020 | ···················· | ···················· | ···················· | ···················· | ···················· | ···················· |
| P | 1273 hCoV-19/Denmark/DCGC-8616/2020\|EPI_ISL_625991\|2020 | ···················· | ···················· | ···················· | ···················· | ···················· | ···················· |
| P | 1274 hCoV-19/Denmark/DCGC-8623/2020\|EPI_ISL_625992\|2020 | ···················· | ···················· | ···················· | ···················· | ···················· | ···················· |
| P | 1275 hCoV-19/Denmark/DCGC-8633/2020\|EPI_ISL_625993\|2020 | ···················· | ···················· | ···················· | ···················· | ···················· | ···················· |
| P | 1276 hCoV-19/Denmark/DCGC-8651/2020\|EPI_ISL_625994\|2020 | ···················· | ···················· | ···················· | ···················· | ···················· | ···················· |
| P | 1277 hCoV-19/Denmark/DCGC-8624/2020\|EPI_ISL_625995\|2020 | ···················· | ···················· | ···················· | ···················· | ···················· | ···················· |
| P | 1278 hCoV-19/Denmark/DCGC-8634/2020\|EPI_ISL_625996\|2020 | ···················· | ···················· | ···················· | ···················· | ···················· | ···················· |
| P | 1279 hCoV-19/Denmark/DCGC-8663/2020\|EPI_ISL_625997\|2020 | ···················· | ···················· | ···················· | ···················· | ···················· | ···················· |
| P | 1280 hCoV-19/Denmark/DCGC-8617/2020\|EPI_ISL_625999\|2020 | ···················· | ···················· | ···················· | ···················· | ···················· | ···················· |
| P | 1281 hCoV-19/Denmark/DCGC-3197/2020\|EPI_ISL_616020\|2020 | ···················· | ···················· | ···················· | ···················· | ···················· | ···················· |
| P | 1282 hCoV-19/Denmark/DCGC-3198/2020\|EPI_ISL_616021\|2020 | ···················· | ···················· | ···················· | ···················· | ···················· | ···················· |
| P | 1283 hCoV-19/Denmark/DCGC-3199/2020\|EPI_ISL_616022\|2020 | ···················· | ···················· | ···················· | ···················· | ···················· | ···················· |
| P | 1284 hCoV-19/Denmark/DCGC-3202/2020\|EPI_ISL_616025\|2020 | ···················· | ···················· | ···················· | ···················· | ···················· | ···················· |
| P | 1285 hCoV-19/Denmark/DCGC-3203/2020\|EPI_ISL_616026\|2020 | ···················· | ···················· | ···················· | ···················· | ···················· | ···················· |
| P | 1286 hCoV-19/Denmark/DCGC-3204/2020\|EPI_ISL_616027\|2020 | ···················· | ···················· | ···················· | ···················· | ···················· | ···················· |
| P | 1287 hCoV-19/Denmark/DCGC-3205/2020\|EPI_ISL_616028\|2020 | ···················· | ···················· | ···················· | ···················· | ···················· | ···················· |
| P | 1288 hCoV-19/Denmark/DCGC-3206/2020\|EPI_ISL_616029\|2020 | ···················· | ···················· | ···················· | ···················· | ···················· | ···················· |
| P | 1289 hCoV-19/Denmark/DCGC-3306/2020\|EPI_ISL_616060\|2020 | ···················· | ···················· | ···················· | ···················· | ···················· | ···················· |
| P | 1290 hCoV-19/Denmark/DCGC-3307/2020\|EPI_ISL_616061\|2020 | ···················· | ···················· | ···················· | ···················· | ···················· | ···················· |
| P | 1291 hCoV-19/Denmark/DCGC-3308/2020\|EPI_ISL_616062\|2020 | ···················· | ····G··············· | ···················· | ···················· | ···················· | ···················· |
| P | 1292 hCoV-19/Denmark/DCGC-3309/2020\|EPI_ISL_616063\|2020 | ···················· | ···················· | ···················· | ···················· | ···················· | ···················· |
| P | 1293 hCoV-19/Denmark/DCGC-3310/2020\|EPI_ISL_616064\|2020 | ···················· | ···················· | ···················· | ···················· | ···················· | ···················· |
| P | 1294 hCoV-19/Denmark/DCGC-3312/2020\|EPI_ISL_616066\|2020 | ···················· | ·A·················· | ···················· | ···················· | ···················· | ···A················ |
| P | 1295 hCoV-19/Denmark/DCGC-3314/2020\|EPI_ISL_616068\|2020 | ···················· | ···················· | ···················· | ···················· | ···················· | ···················· |
| P | 1296 hCoV-19/Denmark/DCGC-3315/2020\|EPI_ISL_616069\|2020 | ···················· | ···················· | ···················· | ···················· | ···················· | ···················· |
| P | 1297 hCoV-19/Denmark/DCGC-3316/2020\|EPI_ISL_616070\|2020 | ···················· | ···················· | ···················· | ···················· | ···················· | ···················· |
| P | 1298 hCoV-19/Denmark/DCGC-3317/2020\|EPI_ISL_616071\|2020 | ···················· | ···················· | ···················· | ···················· | ···················· | ···················· |
| P | 1299 hCoV-19/Denmark/DCGC-3318/2020\|EPI_ISL_616072\|2020 | ···················· | ···················· | ···················· | ···················· | ···················· | ···················· |
| P | 1300 hCoV-19/Denmark/DCGC-3319/2020\|EPI_ISL_616073\|2020 | ···················· | ···················· | ···················· | ···················· | ···················· | ···················· |
| P | 1301 hCoV-19/Denmark/DCGC-3320/2020\|EPI_ISL_616074\|2020 | ···················· | ···················· | ···················· | ···················· | ···················· | ···················· |
| P | 1302 hCoV-19/Denmark/DCGC-3321/2020\|EPI_ISL_616075\|2020 | ···················· | ···················· | ···················· | ···················· | ···················· | ···················· |
| P | 1303 hCoV-19/Denmark/DCGC-3322/2020\|EPI_ISL_616076\|2020 | ···················· | ···················· | ···················· | ···················· | ···················· | ···················· |
| P | 1304 hCoV-19/Denmark/DCGC-3323/2020\|EPI_ISL_616077\|2020 | ···················· | ···················· | ···················· | ···················· | ···················· | ···················· |
| P | 1305 hCoV-19/Denmark/DCGC-3324/2020\|EPI_ISL_616078\|2020 | ···················· | ···················· | ···················· | ···················· | ···················· | ···················· |
| P | 1306 hCoV-19/Denmark/DCGC-3325/2020\|EPI_ISL_616079\|2020 | ···················· | ···················· | ···················· | ···················· | ···················· | ···················· |
| P | 1307 hCoV-19/Denmark/DCGC-3326/2020\|EPI_ISL_616080\|2020 | ···················· | ···················· | ···················· | ···················· | ···················· | ···················· |
| P | 1308 hCoV-19/Denmark/DCGC-3327/2020\|EPI_ISL_616081\|2020 | ···················· | ····G··············· | ···················· | ···················· | ···················· | ···················· |
| P | 1309 hCoV-19/Denmark/DCGC-3328/2020\|EPI_ISL_616082\|2020 | ···················· | ····G··············· | ···················· | ···················· | ···················· | ···················· |
| P | 1310 hCoV-19/Denmark/DCGC-3329/2020\|EPI_ISL_616083\|2020 | ···················· | ···················· | ···················· | ···················· | ···················· | ···················· |
| P | 1311 hCoV-19/Denmark/DCGC-3330/2020\|EPI_ISL_616084\|2020 | ···················· | ···················· | ···················· | ···················· | ···················· | ···················· |
| P | 1312 hCoV-19/Denmark/DCGC-3331/2020\|EPI_ISL_616085\|2020 | ···················· | ···················· | ···················· | ···················· | ···················· | ···················· |
| P | 1313 hCoV-19/Denmark/DCGC-3332/2020\|EPI_ISL_616086\|2020 | ···················· | ···················· | ···················· | ···················· | ···················· | ···················· |
| P | 1314 hCoV-19/Denmark/DCGC-3333/2020\|EPI_ISL_616087\|2020 | ···················· | ···················· | ···················· | ···················· | ···················· | ···················· |

```
P    1315 hCoV-19/Denmark/DCGC-3334/2020|EPI_ISL_616088|2020   ····················  ····················  ·····················  ····················  ····················  ····················
P    1316 hCoV-19/Denmark/DCGC-3335/2020|EPI_ISL_616089|2020   ····················  ····················  ·····················  ····················  ····················  ····················
P    1317 hCoV-19/Denmark/DCGC-3336/2020|EPI_ISL_616090|2020   ····················  ····················  ·····················  ····················  ····················  ····················
P    1318 hCoV-19/Denmark/DCGC-3337/2020|EPI_ISL_616091|2020   ····················  ····················  ·····················  ····················  ····················  ····················
P    1319 hCoV-19/Denmark/DCGC-3338/2020|EPI_ISL_616092|2020   ····················  ····················  ·····················  ····················  ····················  ····················
P    1320 hCoV-19/Denmark/DCGC-3339/2020|EPI_ISL_616093|2020   ····················  ····················  ·····················  ····················  ····················  ····················
P    1321 hCoV-19/Denmark/DCGC-3340/2020|EPI_ISL_616094|2020   ····················  ····················  ·····················  ····················  ····················  ····················
P    1322 hCoV-19/Denmark/DCGC-3341/2020|EPI_ISL_616095|2020   ····················  ····················  ·····················  ····················  ····················  ····················
P    1323 hCoV-19/Denmark/DCGC-3342/2020|EPI_ISL_616096|2020   ····················  ····················  ·····················  ····················  ····················  ····················
P    1324 hCoV-19/Denmark/DCGC-3343/2020|EPI_ISL_616097|2020   ····················  ····················  ·····················  ····················  ····················  ····················
P    1325 hCoV-19/Denmark/DCGC-3345/2020|EPI_ISL_616099|2020   ····················  ····················  ·····················  ····················  ····················  ····················
P    1326 hCoV-19/Denmark/DCGC-3346/2020|EPI_ISL_616100|2020   ····················  ····················  ·····················  ····················  ····················  ····················
P    1327 hCoV-19/Denmark/DCGC-3347/2020|EPI_ISL_616101|2020   ····················  ····················  ·····················  ····················  ····················  ····················
P    1328 hCoV-19/Denmark/DCGC-3348/2020|EPI_ISL_616102|2020   ····················  ····················  ·····················  ····················  ····················  ····················
P    1329 hCoV-19/Denmark/DCGC-3349/2020|EPI_ISL_616103|2020   ····················  ····················  ·····················  ····················  ····················  ····················
P    1330 hCoV-19/Denmark/DCGC-3350/2020|EPI_ISL_616104|2020   ····················  ····················  ·····················  ····················  ····················  ····················
P    1331 hCoV-19/Denmark/DCGC-3351/2020|EPI_ISL_616105|2020   ····················  ····················  ·····················  ····················  ····················  ····················
P    1332 hCoV-19/Denmark/DCGC-3352/2020|EPI_ISL_616106|2020   ········T···········  ····················  ·····················  ····················  ····················  ····················
P    1333 hCoV-19/Denmark/DCGC-3354/2020|EPI_ISL_616108|2020   ····················  ····················  ·····················  ····················  ····················  ····················
P    1334 hCoV-19/Denmark/DCGC-3259/2020|EPI_ISL_616109|2020   ····················  ····················  ·····················  ····················  ····················  ····················
P    1335 hCoV-19/Denmark/DCGC-3260/2020|EPI_ISL_616110|2020   ····················  ····················  ·····················  ····················  ····················  ····················
P    1336 hCoV-19/Denmark/DCGC-3261/2020|EPI_ISL_616111|2020   ····················  ····················  ·····················  ····················  ····················  ····················
P    1337 hCoV-19/Denmark/DCGC-3355/2020|EPI_ISL_616112|2020   ····················  ····················  ·····················  ····················  ····················  ····················
P    1338 hCoV-19/Denmark/DCGC-3356/2020|EPI_ISL_616113|2020   ····················  ····················  ·····················  ····················  ····················  ····················
P    1339 hCoV-19/Denmark/DCGC-3357/2020|EPI_ISL_616114|2020   ····················  ·A··················  ·····················  ····················  ····················  ····················
P    1340 hCoV-19/Denmark/DCGC-3358/2020|EPI_ISL_616115|2020   ····················  ····················  ·····················  ····················  ····················  ····················
P    1341 hCoV-19/Denmark/DCGC-3359/2020|EPI_ISL_616116|2020   ····················  ····················  ·····················  ····················  ····················  ····················
P    1342 hCoV-19/Denmark/DCGC-3360/2020|EPI_ISL_616117|2020   ····················  ····················  ·····················  ····················  ····················  ····················
P    1343 hCoV-19/Denmark/DCGC-3361/2020|EPI_ISL_616118|2020   ····················  ····················  ·····················  ····················  ····················  ····················
P    1344 hCoV-19/Denmark/DCGC-3362/2020|EPI_ISL_616119|2020   ····················  ····················  ·····················  ····················  ····················  ····················
P    1345 hCoV-19/Denmark/DCGC-3363/2020|EPI_ISL_616120|2020   ····················  ····················  ·····················  ····················  ····················  ····················
P    1346 hCoV-19/Denmark/DCGC-3364/2020|EPI_ISL_616121|2020   ····················  ····················  ·····················  ····················  ····················  ····················
P    1347 hCoV-19/Denmark/DCGC-3365/2020|EPI_ISL_616122|2020   ····················  ····················  ·····················  ····················  ····················  ····················
P    1348 hCoV-19/Denmark/DCGC-3366/2020|EPI_ISL_616123|2020   ····················  ····················  ·····················  ····················  ····················  ····················
P    1349 hCoV-19/Denmark/DCGC-3367/2020|EPI_ISL_616124|2020   ····················  ····················  ·····················  ····················  ····················  ····················
P    1350 hCoV-19/Denmark/DCGC-3368/2020|EPI_ISL_616125|2020   ····················  ····················  ·····················  ····················  ····················  ····················
P    1351 hCoV-19/Denmark/DCGC-3369/2020|EPI_ISL_616126|2020   ····················  ····················  ·····················  ····················  ····················  ····················
P    1352 hCoV-19/Denmark/DCGC-3370/2020|EPI_ISL_616127|2020   ····················  ····················  ·····················  ····················  ····················  ····················
P    1353 hCoV-19/Denmark/DCGC-3371/2020|EPI_ISL_616128|2020   ····················  ····················  ·····················  ····················  ····················  ····················
P    1354 hCoV-19/Denmark/DCGC-3372/2020|EPI_ISL_616129|2020   ····················  ····················  ·····················  ····················  ····················  ····················
P    1355 hCoV-19/Denmark/DCGC-3262/2020|EPI_ISL_616130|2020   ····················  ····················  ·····················  ····················  ····················  ····················
P    1356 hCoV-19/Denmark/DCGC-3373/2020|EPI_ISL_616131|2020   ····················  ····················  ·····················  ····················  ····················  ····················
P    1357 hCoV-19/Denmark/DCGC-3374/2020|EPI_ISL_616132|2020   ····················  ····················  ·····················  ····················  ····················  ····················
P    1358 hCoV-19/Denmark/DCGC-3263/2020|EPI_ISL_616133|2020   ····················  ·A··················  ·····················  ····················  ····················  ···A················
P    1359 hCoV-19/Denmark/DCGC-3375/2020|EPI_ISL_616134|2020   ····················  ····················  ·····················  ····················  ····················  ····················
P    1360 hCoV-19/Denmark/DCGC-3376/2020|EPI_ISL_616135|2020   ····················  ····················  ·····················  ····················  ····················  ····················
P    1361 hCoV-19/Denmark/DCGC-3377/2020|EPI_ISL_616136|2020   ····················  ····················  ·····················  ····················  ····················  ····················
P    1362 hCoV-19/Denmark/DCGC-3378/2020|EPI_ISL_616137|2020   ····················  ····················  ·····················  ····················  ····················  ····················
P    1363 hCoV-19/Denmark/DCGC-3379/2020|EPI_ISL_616138|2020   ····················  ····················  ·····················  ····················  ····················  ····················
P    1364 hCoV-19/Denmark/DCGC-3380/2020|EPI_ISL_616139|2020   ····················  ·A··················  ·····················  ····················  ····················  ···A················
P    1365 hCoV-19/Denmark/DCGC-8371/2020|EPI_ISL_617350|2020   ····················  ····················  ·····················  ····················  ····················  ····················
P    1366 hCoV-19/Denmark/DCGC-8388/2020|EPI_ISL_617351|2020   ····················  ····················  ·····················  ····················  ····················  ····················
P    1367 hCoV-19/Denmark/DCGC-8403/2020|EPI_ISL_617352|2020   ····················  ····················  ·····················  ····················  ····················  ····················
P    1368 hCoV-19/Denmark/DCGC-8349/2020|EPI_ISL_617354|2020   ····················  ····················  ·····················  ····················  ····················  ····················
P    1369 hCoV-19/Denmark/DCGC-8360/2020|EPI_ISL_617355|2020   ····················  ····················  ·····················  ····················  ····················  ····················
P    1370 hCoV-19/Denmark/DCGC-8372/2020|EPI_ISL_617356|2020   ····················  ·A··················  ·····················  ····················  ····················  ····················
P    1371 hCoV-19/Denmark/DCGC-8378/2020|EPI_ISL_617357|2020   ····················  ·A··················  ·····················  ····················  ····················  ····················
P    1372 hCoV-19/Denmark/DCGC-8396/2020|EPI_ISL_617358|2020   ····················  ····················  ·····················  ····················  ····················  ····················
P    1373 hCoV-19/Denmark/DCGC-8404/2020|EPI_ISL_617359|2020   ····················  ····················  ·····················  ····················  ····················  ····················
P    1374 hCoV-19/Denmark/DCGC-8473/2020|EPI_ISL_617390|2020   ····················  ····················  ·····················  ····················  ····················  ····················
P    1375 hCoV-19/Denmark/DCGC-8410/2020|EPI_ISL_617391|2020   ····················  ···A················  ·····················  ····················  ····················  ····················
P    1376 hCoV-19/Denmark/DCGC-8417/2020|EPI_ISL_617392|2020   ····················  ····················  ·····················  ····················  ····················  ····················
P    1377 hCoV-19/Denmark/DCGC-8422/2020|EPI_ISL_617393|2020   ····················  ····················  ·····················  ····················  ····················  ····················
P    1378 hCoV-19/Denmark/DCGC-8442/2020|EPI_ISL_617394|2020   ····················  ····················  ·····················  ····················  ····················  ····················
P    1379 hCoV-19/Denmark/DCGC-8455/2020|EPI_ISL_617395|2020   ····················  ·A··················  ·····················  ····················  ····················  ···A················
P    1380 hCoV-19/Denmark/DCGC-8411/2020|EPI_ISL_617396|2020   ····················  ····················  ·····················  ····················  ····················  ····················
P    1381 hCoV-19/Denmark/DCGC-8418/2020|EPI_ISL_617397|2020   ····················  ·A··················  ·····················  ····················  ····················  ····················
P    1382 hCoV-19/Denmark/DCGC-8448/2020|EPI_ISL_617398|2020   ····················  ····················  ·····················  ····················  ····················  ····················
P    1383 hCoV-19/Denmark/DCGC-8456/2020|EPI_ISL_617399|2020   ····················  ····················  ·····················  ····················  ····················  ····················
P    1384 hCoV-19/Denmark/DCGC-8468/2020|EPI_ISL_617400|2020   ····················  ····················  ·····················  ····················  ····················  ····················
P    1385 hCoV-19/Denmark/DCGC-8475/2020|EPI_ISL_617401|2020   ····················  ····················  ·····················  ····················  ····················  ····················
P    1386 hCoV-19/Denmark/DCGC-8424/2020|EPI_ISL_617402|2020   ····················  ····················  ·····················  ····················  ····················  ····················
P    1387 hCoV-19/Denmark/DCGC-8436/2020|EPI_ISL_617403|2020   ····················  ····················  ·····················  ····················  ····················  ····················
P    1388 hCoV-19/Denmark/DCGC-8449/2020|EPI_ISL_617404|2020   ····················  ····················  ·····················  ····················  ····················  ····················
P    1389 hCoV-19/Denmark/DCGC-8457/2020|EPI_ISL_617405|2020   ····················  ····················  ·····················  ····················  ····················  ····················
P    1390 hCoV-19/Denmark/DCGC-2446/2020|EPI_ISL_617406|2020   ····················  ····················  ·····················  ····················  ····················  ····················
P    1391 hCoV-19/Denmark/DCGC-2447/2020|EPI_ISL_617407|2020   ····················  ····················  ·····················  ····················  ····················  ····················
P    1392 hCoV-19/Denmark/DCGC-2448/2020|EPI_ISL_617408|2020   ····················  ····················  ·····················  ····················  ····················  ····················
P    1393 hCoV-19/Denmark/DCGC-2449/2020|EPI_ISL_617409|2020   ····················  ····················  ·····················  ····················  ····················  ····················
P    1394 hCoV-19/Denmark/DCGC-2450/2020|EPI_ISL_617410|2020   ····················  ····················  ·····················  ····················  ····················  ····················
```

| | | | | | | | | |
|---|---|---|---|---|---|---|---|---|
| P | 1395 | hCoV-19/Denmark/DCGC-2451/2020\|EPI_ISL_617411\|2020 | ···················· | ···················· | ···················· | ···················· | ···················· | ···················· |
| P | 1396 | hCoV-19/Denmark/DCGC-2452/2020\|EPI_ISL_617412\|2020 | ···················· | ···················· | ···················· | ···················· | ···················· | ···················· |
| P | 1397 | hCoV-19/Denmark/DCGC-2453/2020\|EPI_ISL_617413\|2020 | ···················· | ···················· | ···················· | ···················· | ···················· | ···················· |
| P | 1398 | hCoV-19/Denmark/DCGC-2454/2020\|EPI_ISL_617414\|2020 | ···················· | ···················· | ···················· | ···················· | ···················· | ···················· |
| P | 1399 | hCoV-19/Denmark/DCGC-2456/2020\|EPI_ISL_617416\|2020 | ···················· | ·A·················· | ···················· | ···················· | ···················· | ···················· |
| P | 1400 | hCoV-19/Denmark/DCGC-2457/2020\|EPI_ISL_617417\|2020 | ···················· | ·A·················· | ···················· | ···················· | ···················· | ···················· |
| P | 1401 | hCoV-19/Denmark/DCGC-2459/2020\|EPI_ISL_617419\|2020 | ···················· | ···················· | ···················· | ···················· | ···················· | ···················· |
| P | 1402 | hCoV-19/Denmark/DCGC-2460/2020\|EPI_ISL_617420\|2020 | ···················· | ···················· | ···················· | ···················· | ···················· | ···················· |
| P | 1403 | hCoV-19/Denmark/DCGC-2462/2020\|EPI_ISL_617422\|2020 | ···················· | ···················· | ···················· | ···················· | ···················· | ···················· |
| P | 1404 | hCoV-19/Denmark/DCGC-2463/2020\|EPI_ISL_617424\|2020 | ···················· | ···················· | ···················· | ···················· | ···················· | ···················· |
| P | 1405 | hCoV-19/Denmark/DCGC-2464/2020\|EPI_ISL_617425\|2020 | ···················· | ···················· | ···················· | ···················· | ···················· | ···················· |
| P | 1406 | hCoV-19/Denmark/DCGC-2465/2020\|EPI_ISL_617426\|2020 | ···················· | ···················· | ···················· | ···················· | ···················· | ···················· |
| P | 1407 | hCoV-19/Denmark/DCGC-2466/2020\|EPI_ISL_617427\|2020 | ···················· | ·A·················· | ···················· | ···················· | ···················· | ···················· |
| P | 1408 | hCoV-19/Denmark/DCGC-2467/2020\|EPI_ISL_617428\|2020 | ···················· | ···················· | ···················· | ···················· | ···················· | ···················· |
| P | 1409 | hCoV-19/Denmark/DCGC-2468/2020\|EPI_ISL_617429\|2020 | ···················· | ···················· | ···················· | ···················· | ···················· | ···················· |
| P | 1410 | hCoV-19/Denmark/DCGC-2250/2020\|EPI_ISL_617430\|2020 | ···················· | ···················· | ···················· | ···················· | ···················· | ···················· |
| P | 1411 | hCoV-19/Denmark/DCGC-2251/2020\|EPI_ISL_617431\|2020 | ···················· | ···················· | ···················· | ···················· | ···················· | ···················· |
| P | 1412 | hCoV-19/Denmark/DCGC-2252/2020\|EPI_ISL_617432\|2020 | ···················· | ···················· | ···················· | ···················· | ···················· | ···················· |
| P | 1413 | hCoV-19/Denmark/DCGC-2253/2020\|EPI_ISL_617433\|2020 | ···················· | ···················· | ···················· | ···················· | ···················· | ···················· |
| P | 1414 | hCoV-19/Denmark/DCGC-2254/2020\|EPI_ISL_617434\|2020 | ···················· | ···················· | ···················· | ···················· | ···················· | ···················· |
| P | 1415 | hCoV-19/Denmark/DCGC-2255/2020\|EPI_ISL_617435\|2020 | ···················· | ·A·················· | ···················· | ···················· | ···················· | ···················· |
| P | 1416 | hCoV-19/Denmark/DCGC-2256/2020\|EPI_ISL_617436\|2020 | ···················· | ···················· | ···················· | ···················· | ···················· | ···················· |
| P | 1417 | hCoV-19/Denmark/DCGC-2257/2020\|EPI_ISL_617437\|2020 | ···················· | ···················· | ···················· | ···················· | ···················· | ···················· |
| P | 1418 | hCoV-19/Denmark/DCGC-2258/2020\|EPI_ISL_617438\|2020 | ···················· | ···················· | ···················· | ···················· | ···················· | ···················· |
| P | 1419 | hCoV-19/Denmark/DCGC-2259/2020\|EPI_ISL_617439\|2020 | ···················· | ···················· | ···················· | ···················· | ···················· | ···················· |
| P | 1420 | hCoV-19/Denmark/DCGC-2260/2020\|EPI_ISL_617440\|2020 | ···················· | ···················· | ···················· | ···················· | ···················· | ···················· |
| P | 1421 | hCoV-19/Denmark/DCGC-2143/2020\|EPI_ISL_617441\|2020 | ···················· | ···················· | ···················· | ···················· | ···················· | ···················· |
| P | 1422 | hCoV-19/Denmark/DCGC-2145/2020\|EPI_ISL_617443\|2020 | ···················· | ···················· | ···················· | ···················· | ···················· | ···················· |
| P | 1423 | hCoV-19/Denmark/DCGC-2148/2020\|EPI_ISL_617446\|2020 | ···················· | ···················· | ···················· | ···················· | ···················· | ···················· |
| P | 1424 | hCoV-19/Denmark/DCGC-2149/2020\|EPI_ISL_617447\|2020 | ···················· | ···················· | ···················· | ···················· | ···················· | ···················· |
| P | 1425 | hCoV-19/Denmark/DCGC-2150/2020\|EPI_ISL_617448\|2020 | ···················· | ···················· | ···················· | ···················· | ···················· | ···················· |
| P | 1426 | hCoV-19/Denmark/DCGC-2151/2020\|EPI_ISL_617449\|2020 | ···················· | ···················· | ···················· | ···················· | ···················· | ···················· |
| P | 1427 | hCoV-19/Denmark/DCGC-2152/2020\|EPI_ISL_617450\|2020 | ···················· | ···················· | ···················· | ···················· | ···················· | ···················· |
| P | 1428 | hCoV-19/Denmark/DCGC-2153/2020\|EPI_ISL_617451\|2020 | ···················· | ···················· | ···················· | ···················· | ···················· | ···················· |
| P | 1429 | hCoV-19/Denmark/DCGC-2154/2020\|EPI_ISL_617452\|2020 | ···················· | ···················· | ···················· | ···················· | ···················· | ···················· |
| P | 1430 | hCoV-19/Denmark/DCGC-2155/2020\|EPI_ISL_617453\|2020 | ···················· | ···················· | ···················· | ···················· | ···················· | ···················· |
| P | 1431 | hCoV-19/Denmark/DCGC-2422/2020\|EPI_ISL_617454\|2020 | ···················· | ···················· | ···················· | ···················· | ···················· | ···················· |
| P | 1432 | hCoV-19/Denmark/DCGC-2156/2020\|EPI_ISL_617455\|2020 | ···················· | ···················· | ···················· | ···················· | ···················· | ···················· |
| P | 1433 | hCoV-19/Denmark/DCGC-2157/2020\|EPI_ISL_617456\|2020 | ···················· | ···················· | ···················· | ···················· | ···················· | ···················· |
| P | 1434 | hCoV-19/Denmark/DCGC-2159/2020\|EPI_ISL_617458\|2020 | ···················· | ···················· | ···················· | ···················· | ···················· | ···················· |
| P | 1435 | hCoV-19/Denmark/DCGC-2160/2020\|EPI_ISL_617459\|2020 | ···················· | ···················· | ···················· | ···················· | ···················· | ···················· |
| P | 1436 | hCoV-19/Denmark/DCGC-2161/2020\|EPI_ISL_617460\|2020 | ···················· | ···················· | ···················· | ···················· | ···················· | ···················· |
| P | 1437 | hCoV-19/Denmark/DCGC-2162/2020\|EPI_ISL_617461\|2020 | ···················· | ···················· | ···················· | ···················· | ···················· | ···················· |
| P | 1438 | hCoV-19/Denmark/DCGC-2163/2020\|EPI_ISL_617462\|2020 | ···················· | ···················· | ···················· | ···················· | ···················· | ···················· |
| P | 1439 | hCoV-19/Denmark/DCGC-2164/2020\|EPI_ISL_617463\|2020 | ···················· | ···················· | ···················· | ···················· | ···················· | ···················· |
| P | 1440 | hCoV-19/Denmark/DCGC-2165/2020\|EPI_ISL_617464\|2020 | ···················· | ·A·················· | ···················· | ···················· | ···················· | ···················· |
| P | 1441 | hCoV-19/Denmark/DCGC-2166/2020\|EPI_ISL_617465\|2020 | ···················· | ···················· | ···················· | ···················· | ···················· | ···················· |
| P | 1442 | hCoV-19/Denmark/DCGC-2168/2020\|EPI_ISL_617467\|2020 | ···················· | ·A·················· | ···················· | ···················· | ···················· | ···················· |
| P | 1443 | hCoV-19/Denmark/DCGC-2170/2020\|EPI_ISL_617469\|2020 | ···················· | ···················· | ···················· | ···················· | ···················· | ···················· |
| P | 1444 | hCoV-19/Denmark/DCGC-7789/2020\|EPI_ISL_618682\|2020 | ···················· | ·A·················· | ···················· | ···················· | ···················· | ···················· |
| P | 1445 | hCoV-19/Denmark/DCGC-7723/2020\|EPI_ISL_618683\|2020 | ···················· | ···················· | ···················· | ···················· | ···················· | ···················· |
| P | 1446 | hCoV-19/Denmark/DCGC-7753/2020\|EPI_ISL_618685\|2020 | ···················· | ···················· | ···················· | ···················· | ···················· | ···················· |
| P | 1447 | hCoV-19/Denmark/DCGC-7760/2020\|EPI_ISL_618686\|2020 | ···················· | ···················· | ···················· | ···················· | ···················· | ···················· |
| P | 1448 | hCoV-19/Denmark/DCGC-7782/2020\|EPI_ISL_618688\|2020 | ···················· | ···················· | ···················· | ···················· | ···················· | ···················· |
| P | 1449 | hCoV-19/Denmark/DCGC-7790/2020\|EPI_ISL_618689\|2020 | ···················· | ···················· | ···················· | ···················· | ···················· | ···················· |
| P | 1450 | hCoV-19/Denmark/DCGC-7756/2020\|EPI_ISL_618719\|2020 | ···················· | ···················· | ···················· | ···················· | ···················· | ···················· |
| P | 1451 | hCoV-19/Denmark/DCGC-1340/2020\|EPI_ISL_618720\|2020 | ···················· | ···················· | ···················· | ···················· | ···················· | ···················· |
| P | 1452 | hCoV-19/Denmark/DCGC-7769/2020\|EPI_ISL_618722\|2020 | ···················· | ···················· | ···················· | ···················· | ···················· | ···················· |
| P | 1453 | hCoV-19/Denmark/DCGC-7728/2020\|EPI_ISL_618725\|2020 | ·······C············ | ···················· | ···················· | ···················· | ···················· | ···················· |
| P | 1454 | hCoV-19/Denmark/DCGC-7732/2020\|EPI_ISL_618726\|2020 | ···················· | ·A·················· | ···················· | ···················· | ···················· | ···A················ |
| P | 1455 | hCoV-19/Denmark/DCGC-7744/2020\|EPI_ISL_618728\|2020 | ···················· | ···················· | ···················· | ···················· | ···················· | ···················· |
| P | 1456 | hCoV-19/Denmark/DCGC-7750/2020\|EPI_ISL_618729\|2020 | ···················· | ·A·················· | ···················· | ···················· | ···················· | ···················· |
| P | 1457 | hCoV-19/Denmark/DCGC-1341/2020\|EPI_ISL_618730\|2020 | ···················· | ···················· | ···················· | ···················· | ···················· | ···················· |
| P | 1458 | hCoV-19/Denmark/DCGC-7757/2020\|EPI_ISL_618731\|2020 | ···················· | ···················· | ···················· | ···················· | ···················· | ···················· |
| P | 1459 | hCoV-19/Denmark/DCGC-7764/2020\|EPI_ISL_618732\|2020 | ···················· | ···················· | ···················· | ···················· | ···················· | ···················· |
| P | 1460 | hCoV-19/Denmark/DCGC-7770/2020\|EPI_ISL_618733\|2020 | ···················· | ···················· | ···················· | ···················· | ···················· | ···················· |
| P | 1461 | hCoV-19/Denmark/DCGC-7775/2020\|EPI_ISL_618734\|2020 | ···················· | ···················· | ···················· | ···················· | ···················· | ···················· |
| P | 1462 | hCoV-19/Denmark/DCGC-7779/2020\|EPI_ISL_618735\|2020 | ···················· | ···················· | ···················· | ···················· | ···················· | ···················· |
| P | 1463 | hCoV-19/Denmark/DCGC-7787/2020\|EPI_ISL_618736\|2020 | ···················· | ···················· | ···················· | ···················· | ···················· | ···················· |
| P | 1464 | hCoV-19/Denmark/DCGC-7793/2020\|EPI_ISL_618737\|2020 | ···················· | ·A·················· | ···················· | ···················· | ···················· | ···A················ |
| P | 1465 | hCoV-19/Denmark/DCGC-7799/2020\|EPI_ISL_618738\|2020 | ···················· | ···················· | ···················· | ···················· | ····T··············· | ···················· |
| P | 1466 | hCoV-19/Denmark/DCGC-7810/2020\|EPI_ISL_618739\|2020 | ···················· | ·A·················· | ···················· | ···················· | ···················· | ···················· |
| P | 1467 | hCoV-19/Denmark/DCGC-1342/2020\|EPI_ISL_618740\|2020 | ···················· | ···················· | ···················· | ···················· | ···················· | ···················· |
| P | 1468 | hCoV-19/Denmark/DCGC-7816/2020\|EPI_ISL_618741\|2020 | ···················· | ···················· | ···················· | ···················· | ···················· | ···················· |
| P | 1469 | hCoV-19/Denmark/DCGC-7820/2020\|EPI_ISL_618742\|2020 | ···················· | ···················· | ···················· | ···················· | ···················· | ···················· |
| P | 1470 | hCoV-19/Denmark/DCGC-7839/2020\|EPI_ISL_618744\|2020 | ···················· | ···················· | ···················· | ···················· | ···················· | ···················· |
| P | 1471 | hCoV-19/Denmark/DCGC-7794/2020\|EPI_ISL_618745\|2020 | ···················· | ·A·················· | ···················· | ···················· | ···················· | ···A················ |
| P | 1472 | hCoV-19/Denmark/DCGC-7800/2020\|EPI_ISL_618746\|2020 | ···················· | ···················· | ···················· | ···················· | ···················· | ···················· |
| P | 1473 | hCoV-19/Denmark/DCGC-7804/2020\|EPI_ISL_618747\|2020 | ···················· | ······A············· | ···················· | ···················· | ···················· | ···················· |
| P | 1474 | hCoV-19/Denmark/DCGC-7811/2020\|EPI_ISL_618748\|2020 | ···················· | ···················· | ···················· | ···················· | ···················· | ···················· |

| | | | | | | | | |
|---|---|---|---|---|---|---|---|---|
| P | 1475 | hCoV-19/Denmark/DCGC-7821/2020\|EPI_ISL_618749\|2020 | ···················· | ···················· | ····················· | ···················· | ···················· | ···················· |
| P | 1476 | hCoV-19/Denmark/DCGC-7834/2020\|EPI_ISL_618750\|2020 | ···················· | ···················· | ····················· | ···················· | ···················· | ···················· |
| P | 1477 | hCoV-19/Denmark/DCGC-7849/2020\|EPI_ISL_618752\|2020 | ···················· | ···················· | ····················· | ···················· | ···················· | ···················· |
| P | 1478 | hCoV-19/Denmark/DCGC-7805/2020\|EPI_ISL_618754\|2020 | ···················· | ···················· | ····················· | ···················· | ···················· | ···················· |
| P | 1479 | hCoV-19/Denmark/DCGC-7828/2020\|EPI_ISL_618756\|2020 | ···················· | ···················· | ····················· | ···················· | ···················· | ···················· |
| P | 1480 | hCoV-19/Denmark/DCGC-7835/2020\|EPI_ISL_618757\|2020 | ···················· | ···················· | ····················· | ···················· | ···················· | ···················· |
| P | 1481 | hCoV-19/Denmark/DCGC-7795/2020\|EPI_ISL_618759\|2020 | ···················· | ····A··············· | ····················· | ···················· | ···················· | ···················· |
| P | 1482 | hCoV-19/Denmark/DCGC-7801/2020\|EPI_ISL_618760\|2020 | ···················· | ···················· | ····················· | ···················· | ···················· | ···················· |
| P | 1483 | hCoV-19/Denmark/DCGC-7806/2020\|EPI_ISL_618761\|2020 | ···················· | ···················· | ····················· | ···················· | ···················· | ···················· |
| P | 1484 | hCoV-19/Denmark/DCGC-7812/2020\|EPI_ISL_618762\|2020 | ···················· | ···················· | ····················· | ···················· | ···················· | ···················· |
| P | 1485 | hCoV-19/Denmark/DCGC-7823/2020\|EPI_ISL_618763\|2020 | ···················· | ···················· | ····················· | ···················· | ···················· | ···················· |
| P | 1486 | hCoV-19/Denmark/DCGC-7796/2020\|EPI_ISL_618770\|2020 | ···················· | ···················· | ····················· | ···················· | ···················· | ···················· |
| P | 1487 | hCoV-19/Denmark/DCGC-7813/2020\|EPI_ISL_618772\|2020 | ···················· | ···················· | ····················· | ···················· | ···················· | ···················· |
| P | 1488 | hCoV-19/Denmark/DCGC-7830/2020\|EPI_ISL_618774\|2020 | ···················· | ···················· | ····················· | ···················· | ···················· | ···················· |
| P | 1489 | hCoV-19/Denmark/DCGC-7837/2020\|EPI_ISL_618775\|2020 | ···················· | ···················· | ····················· | ···················· | ···················· | ···················· |
| P | 1490 | hCoV-19/Denmark/DCGC-7841/2020\|EPI_ISL_618776\|2020 | ···················· | ···················· | ····················· | ···················· | ···················· | ···················· |
| P | 1491 | hCoV-19/Denmark/DCGC-7845/2020\|EPI_ISL_618777\|2020 | ···················· | ···················· | ····················· | ···················· | ···················· | ···················· |
| P | 1492 | hCoV-19/Denmark/DCGC-7802/2020\|EPI_ISL_618778\|2020 | ···················· | ···················· | ····················· | ···················· | ···················· | ···················· |
| P | 1493 | hCoV-19/Denmark/DCGC-7817/2020\|EPI_ISL_618779\|2020 | ···················· | ···················· | ····················· | ···················· | ···················· | ···················· |
| P | 1494 | hCoV-19/Denmark/DCGC-7825/2020\|EPI_ISL_618780\|2020 | ···················· | ···················· | ····················· | ···················· | ···················· | ···················· |
| P | 1495 | hCoV-19/Denmark/DCGC-7831/2020\|EPI_ISL_618781\|2020 | ···················· | ···················· | ····················· | ···················· | ···················· | ···················· |
| P | 1496 | hCoV-19/Denmark/DCGC-7852/2020\|EPI_ISL_618783\|2020 | ···················· | ···················· | ····················· | ···················· | ···················· | ···················· |
| P | 1497 | hCoV-19/Denmark/DCGC-7797/2020\|EPI_ISL_618784\|2020 | ···················· | ···················· | ····················· | ···················· | ···················· | ···················· |
| P | 1498 | hCoV-19/Denmark/DCGC-7814/2020\|EPI_ISL_618786\|2020 | ···················· | ···················· | ····················· | ···················· | ···················· | ···················· |
| P | 1499 | hCoV-19/Denmark/DCGC-7818/2020\|EPI_ISL_618787\|2020 | ···················· | ···················· | ····················· | ···················· | ···················· | ···················· |
| P | 1500 | hCoV-19/Denmark/DCGC-7798/2020\|EPI_ISL_618791\|2020 | ···················· | ···················· | ····················· | ···················· | ···················· | ···················· |
| P | 1501 | hCoV-19/Denmark/DCGC-7803/2020\|EPI_ISL_618792\|2020 | ···················· | ···················· | ····················· | ···················· | ···················· | ···················· |
| P | 1502 | hCoV-19/Denmark/DCGC-7809/2020\|EPI_ISL_618793\|2020 | ···················· | ···················· | ····················· | ···················· | ···················· | ···················· |
| P | 1503 | hCoV-19/Denmark/DCGC-7815/2020\|EPI_ISL_618794\|2020 | ···················· | ···················· | ····················· | ···················· | ···················· | ···················· |
| P | 1504 | hCoV-19/Denmark/DCGC-7819/2020\|EPI_ISL_618795\|2020 | ···················· | ···················· | ····················· | ···················· | ···················· | ···················· |
| P | 1505 | hCoV-19/Denmark/DCGC-7826/2020\|EPI_ISL_618796\|2020 | ···················· | ···················· | ····················· | ···················· | ···················· | ···················· |
| P | 1506 | hCoV-19/Denmark/DCGC-7842/2020\|EPI_ISL_618798\|2020 | ···················· | ···················· | ····················· | ···················· | ···················· | ···················· |
| P | 1507 | hCoV-19/England/MILK-AB81DD/2020\|EPI_ISL_623250\|20 | ···················· | ···················· | ····················· | ···················· | ···················· | ···················· |
| P | 1508 | hCoV-19/England/MILK-ABB8D5/2020\|EPI_ISL_623251\|20 | ···················· | ···················· | ····················· | ···················· | ···················· | ···················· |
| P | 1509 | hCoV-19/England/MILK-ABB398/2020\|EPI_ISL_623253\|20 | ···················· | ···················· | ····················· | ···················· | ···················· | ···················· |
| P | 1510 | hCoV-19/England/MILK-AB74B7/2020\|EPI_ISL_623254\|20 | ···················· | ·A·················· | ····················· | ···················· | ···················· | ···················· |
| P | 1511 | hCoV-19/England/MILK-AB7FF2/2020\|EPI_ISL_623255\|20 | ···················· | ···················· | ····················· | ···················· | ···················· | ···················· |
| P | 1512 | hCoV-19/England/CAMC-AAFD20/2020\|EPI_ISL_623256\|20 | ···················· | ···················· | ····················· | ···················· | ···················· | ···················· |
| P | 1513 | hCoV-19/England/MILK-ABB495/2020\|EPI_ISL_623258\|20 | ···················· | ···················· | ····················· | ···················· | ···················· | ···················· |
| P | 1514 | hCoV-19/England/MILK-AB7A97/2020\|EPI_ISL_623261\|20 | ···················· | ···················· | ····················· | ···················· | ···················· | ···················· |
| P | 1515 | hCoV-19/England/MILK-AB7A6A/2020\|EPI_ISL_623263\|20 | ···················· | ···················· | ····················· | ···················· | ···················· | ···················· |
| P | 1516 | hCoV-19/England/CAMC-AAFC23/2020\|EPI_ISL_623268\|20 | ···················· | ···················· | ····················· | ···················· | ···················· | ···················· |
| P | 1517 | hCoV-19/England/MILK-ABB6F9/2020\|EPI_ISL_623269\|20 | ···················· | ···················· | ····················· | ···················· | ···················· | ···················· |
| P | 1518 | hCoV-19/England/ALDP-A957A7/2020\|EPI_ISL_624580\|20 | ···················· | ···················· | ····················· | ···················· | ···················· | ···················· |
| P | 1519 | hCoV-19/England/ALDP-A94EB2/2020\|EPI_ISL_624581\|20 | ···················· | ···················· | ····················· | ···················· | ···················· | ···················· |
| P | 1520 | hCoV-19/England/ALDP-A952E2/2020\|EPI_ISL_624582\|20 | ···················· | ···················· | ····················· | ···················· | ···················· | ···················· |
| P | 1521 | hCoV-19/England/ALDP-A95E48/2020\|EPI_ISL_624583\|20 | ···················· | ···················· | ····················· | ···················· | ···················· | ···················· |
| P | 1522 | hCoV-19/England/ALDP-A94F64/2020\|EPI_ISL_624584\|20 | ···················· | ·A·················· | ····················· | ···················· | ···················· | ···A················ |
| P | 1523 | hCoV-19/England/ALDP-A96041/2020\|EPI_ISL_624585\|20 | ···················· | ···················· | ····················· | ···················· | ···················· | ···················· |
| P | 1524 | hCoV-19/England/ALDP-A9426B/2020\|EPI_ISL_624586\|20 | ···················· | ···················· | ····················· | ···················· | ···················· | ···················· |
| P | 1525 | hCoV-19/England/ALDP-A9630C/2020\|EPI_ISL_624587\|20 | ···················· | ···················· | ····················· | ···················· | ···················· | ···················· |
| P | 1526 | hCoV-19/England/ALDP-A94B06/2020\|EPI_ISL_624588\|20 | ···················· | ···················· | ····················· | ···················· | ···················· | ···················· |
| P | 1527 | hCoV-19/England/ALDP-A95886/2020\|EPI_ISL_624589\|20 | ···················· | ···················· | ····················· | ···················· | ···················· | ···················· |
| P | 1528 | hCoV-19/England/ALDP-A96102/2020\|EPI_ISL_624590\|20 | ···················· | ···················· | ····················· | ···················· | ···················· | ···················· |
| P | 1529 | hCoV-19/England/ALDP-A9468D/2020\|EPI_ISL_624591\|20 | ···················· | ···················· | ····················· | ···················· | ···················· | ···················· |
| P | 1530 | hCoV-19/England/ALDP-A96375/2020\|EPI_ISL_624592\|20 | ···················· | ···················· | ····················· | ···················· | ···················· | ···················· |
| P | 1531 | hCoV-19/England/ALDP-A9522E/2020\|EPI_ISL_624594\|20 | ···················· | ···················· | ····················· | ···················· | ···················· | ···················· |
| P | 1532 | hCoV-19/England/ALDP-A94ED0/2020\|EPI_ISL_624595\|20 | ···················· | ···················· | ····················· | ···················· | ···················· | ···················· |
| P | 1533 | hCoV-19/England/ALDP-A94F91/2020\|EPI_ISL_624599\|20 | ···················· | ···················· | ····················· | ···················· | ···················· | ···················· |
| P | 1534 | hCoV-19/Denmark/DCGC-8780/2020\|EPI_ISL_625800\|2020 | ···················· | ···················· | ····················· | ···················· | ···················· | ···················· |
| P | 1535 | hCoV-19/Denmark/DCGC-8758/2020\|EPI_ISL_625801\|2020 | ···················· | ···················· | ····················· | ···················· | ···················· | ···················· |
| P | 1536 | hCoV-19/Denmark/DCGC-8764/2020\|EPI_ISL_625802\|2020 | ···················· | ···················· | ····················· | ···················· | ···················· | ···················· |
| P | 1537 | hCoV-19/Denmark/DCGC-3200/2020\|EPI_ISL_616023\|2020 | ···················· | ···················· | ····················· | ···················· | ···················· | ···················· |
| P | 1538 | hCoV-19/Denmark/DCGC-3201/2020\|EPI_ISL_616024\|2020 | ···················· | ···················· | ····················· | ···················· | ···················· | ···················· |
| P | 1539 | hCoV-19/Denmark/DCGC-3207/2020\|EPI_ISL_616030\|2020 | ···················· | ···················· | ····················· | ···················· | ···················· | ···················· |
| P | 1540 | hCoV-19/Denmark/DCGC-3254/2020\|EPI_ISL_616031\|2020 | ···················· | ···················· | ····················· | ···················· | ···················· | ···················· |
| P | 1541 | hCoV-19/Denmark/DCGC-3255/2020\|EPI_ISL_616032\|2020 | ···················· | ···················· | ····················· | ···················· | ···················· | ···················· |
| P | 1542 | hCoV-19/Denmark/DCGC-3256/2020\|EPI_ISL_616033\|2020 | ···················· | ···················· | ····················· | ···················· | ···················· | ···················· |
| P | 1543 | hCoV-19/Denmark/DCGC-3283/2020\|EPI_ISL_616035\|2020 | ···················· | ···················· | ····················· | ···················· | ···················· | ···················· |
| P | 1544 | hCoV-19/Denmark/DCGC-3284/2020\|EPI_ISL_616036\|2020 | ···················· | ···················· | ····················· | ···················· | ···················· | ···················· |
| P | 1545 | hCoV-19/Denmark/DCGC-3285/2020\|EPI_ISL_616037\|2020 | ···················· | ···················· | ····················· | ···················· | ···················· | ···················· |
| P | 1546 | hCoV-19/Denmark/DCGC-3286/2020\|EPI_ISL_616038\|2020 | ···················· | ···················· | ····················· | ···················· | ···················· | ···················· |
| P | 1547 | hCoV-19/Denmark/DCGC-3287/2020\|EPI_ISL_616039\|2020 | ···················· | ···················· | ····················· | ···················· | ···················· | ···················· |
| P | 1548 | hCoV-19/Denmark/DCGC-3288/2020\|EPI_ISL_616040\|2020 | ···················· | ···················· | ····················· | ···················· | ···················· | ···················· |
| P | 1549 | hCoV-19/Denmark/DCGC-3289/2020\|EPI_ISL_616041\|2020 | ···················· | ···················· | ····················· | ···················· | ···················· | ···················· |
| P | 1550 | hCoV-19/Denmark/DCGC-3290/2020\|EPI_ISL_616042\|2020 | ···················· | ···················· | ····················· | ···················· | ···················· | ···················· |
| P | 1551 | hCoV-19/Denmark/DCGC-3291/2020\|EPI_ISL_616043\|2020 | ···················· | ···················· | ····················· | ···················· | ···················· | ···················· |
| P | 1552 | hCoV-19/Denmark/DCGC-3292/2020\|EPI_ISL_616044\|2020 | ···················· | ···················· | ····················· | ···················· | ···················· | ···················· |
| P | 1553 | hCoV-19/Denmark/DCGC-3293/2020\|EPI_ISL_616045\|2020 | ···················· | ···················· | ····················· | ···················· | ···················· | ···················· |
| P | 1554 | hCoV-19/Denmark/DCGC-3294/2020\|EPI_ISL_616046\|2020 | ···················· | ···················· | ····················· | ···················· | ···················· | ···················· |

P 1555 hCoV-19/Denmark/DCGC-3295/2020|EPI_ISL_616047|2020 ···················· ···················· ····················· ···················· ···················· ····················
P 1556 hCoV-19/Denmark/DCGC-3296/2020|EPI_ISL_616048|2020 ···················· ···················· ····················· ···················· ···················· ····················
P 1557 hCoV-19/Denmark/DCGC-3298/2020|EPI_ISL_616050|2020 ···················· ···················· ····················· ···················· ···················· ····················
P 1558 hCoV-19/Denmark/DCGC-3299/2020|EPI_ISL_616051|2020 ···················· ···················· ····················· ···················· ···················· ····················
P 1559 hCoV-19/Denmark/DCGC-3300/2020|EPI_ISL_616052|2020 ···················· ···················· ····················· ···················· ···················· ····················
P 1560 hCoV-19/Denmark/DCGC-3301/2020|EPI_ISL_616053|2020 ···················· ···················· ····················· ···················· ···················· ····················
P 1561 hCoV-19/Denmark/DCGC-3302/2020|EPI_ISL_616054|2020 ···················· ···················· ····················· ···················· ···················· ····················
P 1562 hCoV-19/Denmark/DCGC-3303/2020|EPI_ISL_616055|2020 ···················· ···················· ····················· ···················· ···················· ····················
P 1563 hCoV-19/Denmark/DCGC-3257/2020|EPI_ISL_616056|2020 ···················· ···················· ····················· ···················· ···················· ····················
P 1564 hCoV-19/Denmark/DCGC-3304/2020|EPI_ISL_616057|2020 ···················· ···················· ····················· ···················· ···················· ····················
P 1565 hCoV-19/Denmark/DCGC-3258/2020|EPI_ISL_616058|2020 ···················· ···················· ····················· ···················· ···················· ····················
P 1566 hCoV-19/Denmark/DCGC-3305/2020|EPI_ISL_616059|2020 ···················· ···················· ····················· ···················· ···················· ····················
P 1567 hCoV-19/Denmark/DCGC-8337/2020|EPI_ISL_617353|2020 ···················· ···················· ····················· ···················· ···················· ····················
P 1568 hCoV-19/Denmark/DCGC-8338/2020|EPI_ISL_617360|2020 ···················· ···················· ····················· ···················· ···················· ····················
P 1569 hCoV-19/Denmark/DCGC-8350/2020|EPI_ISL_617361|2020 ···················· ···················· ····················· ···················· ···················· ····················
P 1570 hCoV-19/Denmark/DCGC-8361/2020|EPI_ISL_617362|2020 ···················· ···················· ····················· ···················· ···················· ····················
P 1571 hCoV-19/Denmark/DCGC-8373/2020|EPI_ISL_617363|2020 ···················· ·A·················· ····················· ···················· ···················· ····················
P 1572 hCoV-19/Denmark/DCGC-8379/2020|EPI_ISL_617364|2020 ···················· ·A·················· ····················· ···················· ···················· ····················
P 1573 hCoV-19/Denmark/DCGC-8390/2020|EPI_ISL_617365|2020 ···················· ···················· ····················· ···················· ···················· ····················
P 1574 hCoV-19/Denmark/DCGC-8397/2020|EPI_ISL_617366|2020 ···················· ···················· ····················· ···················· ···················· ····················
P 1575 hCoV-19/Denmark/DCGC-8339/2020|EPI_ISL_617367|2020 ···················· ···················· ····················· ···················· ···················· ····················
P 1576 hCoV-19/Denmark/DCGC-8344/2020|EPI_ISL_617368|2020 ···················· ···················· ····················· ···················· ···················· ····················
P 1577 hCoV-19/Denmark/DCGC-8356/2020|EPI_ISL_617369|2020 ···················· ···················· ····················· ···················· ···················· ····················
P 1578 hCoV-19/Denmark/DCGC-8383/2020|EPI_ISL_617370|2020 ···················· ···················· ····················· ···················· ···················· ····················
P 1579 hCoV-19/Denmark/DCGC-8398/2020|EPI_ISL_617371|2020 ···················· ···················· ····················· ···················· ···················· ····················
P 1580 hCoV-19/Denmark/DCGC-8345/2020|EPI_ISL_617372|2020 ···················· ···················· ····················· ···················· ···················· ····················
P 1581 hCoV-19/Denmark/DCGC-8352/2020|EPI_ISL_617373|2020 ···················· ···················· ····················· ···················· ···················· ····················
P 1582 hCoV-19/Denmark/DCGC-8363/2020|EPI_ISL_617374|2020 ···················· ···················· ····················· ···················· ···················· ····················
P 1583 hCoV-19/Denmark/DCGC-8380/2020|EPI_ISL_617375|2020 ·······T············ ···················· ····················· ···················· ···················· ····················
P 1584 hCoV-19/Denmark/DCGC-8384/2020|EPI_ISL_617376|2020 ···················· ···················· ····················· ···················· ···················· ····················
P 1585 hCoV-19/Denmark/DCGC-8392/2020|EPI_ISL_617377|2020 ···················· ···················· ····················· ···················· ···················· ····················
P 1586 hCoV-19/Denmark/DCGC-8407/2020|EPI_ISL_617378|2020 ···················· ···················· ····················· ···················· ·····T·············· ····················
P 1587 hCoV-19/Denmark/DCGC-8405/2020|EPI_ISL_617379|2020 ···················· ···················· ····················· ···················· ···················· ····················
P 1588 hCoV-19/Denmark/DCGC-8408/2020|EPI_ISL_617380|2020 ···················· ···················· ····················· ···················· ···················· ····················
P 1589 hCoV-19/Denmark/DCGC-8406/2020|EPI_ISL_617381|2020 ···················· ···················· ····················· ···················· ···················· ····················
P 1590 hCoV-19/Denmark/DCGC-1349/2020|EPI_ISL_617382|2020 ···················· ···················· ····················· ···················· ···················· ····················
P 1591 hCoV-19/Denmark/DCGC-1676/2020|EPI_ISL_617383|2020 ···················· ···················· ····················· ···················· ···················· ····················
P 1592 hCoV-19/Denmark/DCGC-1725/2020|EPI_ISL_617384|2020 ···················· ···················· ····················· ···················· ···················· ····················
P 1593 hCoV-19/Denmark/DCGC-8416/2020|EPI_ISL_617385|2020 ···················· ···················· ····················· ···················· ···················· ····················
P 1594 hCoV-19/Denmark/DCGC-8441/2020|EPI_ISL_617386|2020 ···················· ···················· ····················· ···················· ···················· ····················
P 1595 hCoV-19/Denmark/DCGC-8454/2020|EPI_ISL_617387|2020 ···················· ···················· ····················· ···················· ···················· ····················
P 1596 hCoV-19/Denmark/DCGC-8462/2020|EPI_ISL_617388|2020 ···················· ·A·················· ····················· ···················· ···················· ····················
P 1597 hCoV-19/Denmark/DCGC-8466/2020|EPI_ISL_617389|2020 ···················· ···················· ····················· ···················· ···················· ····················
P 1598 hCoV-19/Denmark/DCGC-7771/2020|EPI_ISL_618680|2020 ···················· ···················· ····················· ···················· ···················· ····················
P 1599 hCoV-19/Denmark/DCGC-7735/2020|EPI_ISL_618684|2020 ···················· ···················· ····················· ···················· ···················· ····················
P 1600 hCoV-19/Denmark/DCGC-7724/2020|EPI_ISL_618690|2020 ···················· ···················· ····················· ···················· ···················· ····················
P 1601 hCoV-19/Denmark/DCGC-7736/2020|EPI_ISL_618691|2020 ···················· ·A·················· ····················· ···················· ···················· ····················
P 1602 hCoV-19/Denmark/DCGC-7746/2020|EPI_ISL_618692|2020 ···················· ···················· ····················· ···················· ···················· ····················
P 1603 hCoV-19/Denmark/DCGC-7754/2020|EPI_ISL_618693|2020 ···················· ···················· ····················· ···················· ···················· ····················
P 1604 hCoV-19/Denmark/DCGC-7761/2020|EPI_ISL_618694|2020 ···················· ···················· ····················· ···················· ···················· ····················
P 1605 hCoV-19/Denmark/DCGC-7777/2020|EPI_ISL_618696|2020 ···················· ···················· ····················· ···················· ···················· ····················
P 1606 hCoV-19/Denmark/DCGC-7791/2020|EPI_ISL_618698|2020 ···················· ···················· ····················· ···················· ···················· ····················
P 1607 hCoV-19/Denmark/DCGC-7053/2020|EPI_ISL_619902|2020 ···················· ···················· ····················· ···················· ···················· ····················
P 1608 hCoV-19/Denmark/DCGC-7054/2020|EPI_ISL_619903|2020 ···················· ···················· ····················· ···················· ···················· ····················
P 1609 hCoV-19/Denmark/DCGC-1643/2020|EPI_ISL_619905|2020 ···················· ···················· ····················· ···················· ···················· ····················
P 1610 hCoV-19/Denmark/DCGC-5757/2020|EPI_ISL_619906|2020 ···················· ···················· ····················· ···················· ···················· ····················
P 1611 hCoV-19/Denmark/DCGC-5758/2020|EPI_ISL_619907|2020 ···················· ···················· ····················· ···················· ···················· ····················
P 1612 hCoV-19/Denmark/DCGC-5759/2020|EPI_ISL_619908|2020 ···················· ···················· ····················· ···················· ···················· ····················
P 1613 hCoV-19/Denmark/DCGC-5760/2020|EPI_ISL_619909|2020 ···················· ···················· ····················· ···················· ···················· ····················
P 1614 hCoV-19/Denmark/DCGC-5761/2020|EPI_ISL_619910|2020 ···················· ···················· ····················· ···················· ···················· ····················
P 1615 hCoV-19/Denmark/DCGC-5762/2020|EPI_ISL_619911|2020 ···················· ···················· ····················· ···················· ···················· ····················
P 1616 hCoV-19/Denmark/DCGC-5763/2020|EPI_ISL_619912|2020 ···················· ···················· ····················· ···················· ···················· ····················
P 1617 hCoV-19/Denmark/DCGC-5627/2020|EPI_ISL_619914|2020 ···················· ···················· ····················· ···················· ···················· ····················
P 1618 hCoV-19/Denmark/DCGC-5628/2020|EPI_ISL_619915|2020 ···················· ···················· ····················· ···················· ···················· ····················
P 1619 hCoV-19/Denmark/DCGC-5629/2020|EPI_ISL_619916|2020 ···················· ···················· ····················· ···················· ···················· ····················
P 1620 hCoV-19/Denmark/DCGC-5630/2020|EPI_ISL_619917|2020 ···················· ···················· ····················· ···················· ···················· ····················
P 1621 hCoV-19/Denmark/DCGC-5639/2020|EPI_ISL_619926|2020 ···················· ···················· ····················· ···················· ···················· ····················
P 1622 hCoV-19/Denmark/DCGC-5575/2020|EPI_ISL_620900|2020 ···················· ···················· ····················· ···················· ···················· ····················
P 1623 hCoV-19/Denmark/DCGC-5576/2020|EPI_ISL_620901|2020 ···················· ···················· ····················· ···················· ···················· ····················
P 1624 hCoV-19/Denmark/DCGC-5577/2020|EPI_ISL_620902|2020 ···················· ···················· ····················· ···················· ···················· ····················
P 1625 hCoV-19/Denmark/DCGC-5578/2020|EPI_ISL_620903|2020 ···················· ···················· ····················· ···················· ···················· ····················
P 1626 hCoV-19/Denmark/DCGC-5579/2020|EPI_ISL_620904|2020 ···················· ···················· ····················· ···················· ···················· ····················
P 1627 hCoV-19/Denmark/DCGC-5580/2020|EPI_ISL_620905|2020 ···················· ···················· ····················· ···················· ···················· ····················
P 1628 hCoV-19/Denmark/DCGC-5581/2020|EPI_ISL_620906|2020 ···················· ···················· ····················· ···················· ···················· ····················
P 1629 hCoV-19/Denmark/DCGC-5583/2020|EPI_ISL_620908|2020 ···················· ···················· ····················· ···················· ···················· ····················
P 1630 hCoV-19/Denmark/DCGC-5584/2020|EPI_ISL_620909|2020 ···················· ···················· ····················· ···················· ···················· ····················
P 1631 hCoV-19/Denmark/DCGC-5809/2020|EPI_ISL_620996|2020 ···················· ···················· ····················· ···················· ···················· ····················
P 1632 hCoV-19/Denmark/DCGC-5810/2020|EPI_ISL_620997|2020 ···················· ···················· ····················· ···················· ···················· ····················
P 1633 hCoV-19/Denmark/DCGC-5812/2020|EPI_ISL_620999|2020 ···················· ···················· ····················· ···················· ···················· ····················
P 1634 hCoV-19/Brazil/RJ-00349/2020|EPI_ISL_623152|2020-0 ···················· ···················· ····················· ···················· ···················· ····················

| | | | | | | | |
|---|---|---|---|---|---|---|---|
| P | 1635 hCoV-19/Brazil/RJ-00351/2020\|EPI_ISL_623154\|2020-0 | ···················· | ···················· | ····················· | ···················· | ···················· | ···················· |
| P | 1636 hCoV-19/Brazil/RJ-00354/2020\|EPI_ISL_623157\|2020-0 | ···················· | ···················· | ····················· | ···················· | ···················· | ···················· |
| P | 1637 hCoV-19/Brazil/RJ-00355/2020\|EPI_ISL_623158\|2020-0 | ···················· | ···················· | ····················· | ···················· | ···················· | ···················· |
| P | 1638 hCoV-19/Brazil/RJ-00357/2020\|EPI_ISL_623160\|2020-0 | ···················· | ···················· | ····················· | ···················· | ···················· | ···················· |
| P | 1639 hCoV-19/Brazil/RJ-00358/2020\|EPI_ISL_623161\|2020-0 | ···················· | ···················· | ····················· | ···················· | ···················· | ···················· |
| P | 1640 hCoV-19/Brazil/RJ-00359/2020\|EPI_ISL_623162\|2020-0 | ···················· | ···················· | ····················· | ···················· | ···················· | ···················· |
| P | 1641 hCoV-19/Brazil/RJ-00361/2020\|EPI_ISL_623164\|2020-0 | ···················· | ···················· | ····················· | ···················· | ···················· | ···················· |
| P | 1642 hCoV-19/Brazil/RJ-00362/2020\|EPI_ISL_623165\|2020-0 | ···················· | ···················· | ····················· | ···················· | ···················· | ···················· |
| P | 1643 hCoV-19/Brazil/RJ-00363/2020\|EPI_ISL_623166\|2020-0 | ···················· | ···················· | ····················· | ···················· | ···················· | ···················· |
| P | 1644 hCoV-19/Brazil/RJ-00365/2020\|EPI_ISL_623168\|2020-0 | ···················· | ···················· | ····················· | ···················· | ···················· | ···················· |
| P | 1645 hCoV-19/USA/UT-UPHL-201113067/2020\|EPI_ISL_623180\| | ···················· | ···················· | ····················· | ···················· | ···················· | ···················· |
| P | 1646 hCoV-19/USA/UT-UPHL-201121884/2020\|EPI_ISL_623182\| | ···················· | ···················· | ····················· | ···················· | ···················· | ···················· |
| P | 1647 hCoV-19/USA/UT-UPHL-201116971/2020\|EPI_ISL_623183\| | ···················· | ···················· | ····················· | ···················· | ···················· | ···················· |
| P | 1648 hCoV-19/USA/UT-UPHL-201101322/2020\|EPI_ISL_623185\| | ···················· | ···················· | ····················· | ···················· | ···················· | ···················· |
| P | 1649 hCoV-19/USA/UT-UPHL-201108812/2020\|EPI_ISL_623186\| | ···················· | ···················· | ····················· | ···················· | ···················· | ···················· |
| P | 1650 hCoV-19/USA/UT-UPHL-201119264/2020\|EPI_ISL_623188\| | ···················· | ···················· | ····················· | ···················· | ···················· | ···················· |
| P | 1651 hCoV-19/USA/UT-UPHL-201114015/2020\|EPI_ISL_623189\| | ···················· | ···················· | ····················· | ···················· | ···················· | ···················· |
| P | 1652 hCoV-19/USA/UT-UPHL-201103072/2020\|EPI_ISL_623200\| | ···················· | ···················· | ····················· | ···················· | ···················· | ···················· |
| P | 1653 hCoV-19/USA/UT-UPHL-201109982/2020\|EPI_ISL_623202\| | ···················· | ···················· | ····················· | ···················· | ···················· | ···················· |
| P | 1654 hCoV-19/USA/UT-UPHL-201130116/2020\|EPI_ISL_623203\| | ···················· | ···················· | ····················· | ···················· | ···················· | ···················· |
| P | 1655 hCoV-19/USA/UT-UPHL-201104904/2020\|EPI_ISL_623205\| | ···················· | ···················· | ····················· | ···················· | ···················· | ···················· |
| P | 1656 hCoV-19/USA/UT-UPHL-201122369/2020\|EPI_ISL_623207\| | ···················· | ···················· | ····················· | ···················· | ···················· | ···················· |
| P | 1657 hCoV-19/USA/UT-UPHL-201110993/2020\|EPI_ISL_623208\| | ···················· | ···················· | ····················· | ···················· | ···················· | ···················· |
| P | 1658 hCoV-19/England/MILK-AB84D4/2020\|EPI_ISL_623211\|20 | ···················· | ···················· | ····················· | ···················· | ···T················ | ···················· |
| P | 1659 hCoV-19/England/MILK-AB7709/2020\|EPI_ISL_623212\|20 | ···················· | ···················· | ····················· | ···················· | ···················· | ···················· |
| P | 1660 hCoV-19/England/MILK-AB7A3D/2020\|EPI_ISL_623213\|20 | ···················· | ···················· | ····················· | ···················· | ···················· | ···················· |
| P | 1661 hCoV-19/England/MILK-AB86ED/2020\|EPI_ISL_623215\|20 | ···················· | ···················· | ····················· | ···················· | ···················· | ···················· |
| P | 1662 hCoV-19/England/MILK-AB83D7/2020\|EPI_ISL_623216\|20 | ···················· | ·A·················· | ····················· | ···················· | ···················· | ··A················· |
| P | 1663 hCoV-19/England/MILK-ABB92D/2020\|EPI_ISL_623217\|20 | ···················· | ···················· | ····················· | ···················· | ···················· | ···················· |
| P | 1664 hCoV-19/England/MILK-AB7350/2020\|EPI_ISL_623218\|20 | ···················· | ···················· | ····················· | ···················· | ···················· | ···················· |
| P | 1665 hCoV-19/England/MILK-AB8401/2020\|EPI_ISL_623219\|20 | ···················· | ···················· | ····················· | ···················· | ···················· | ···················· |
| P | 1666 hCoV-19/England/CAMC-AAFACF/2020\|EPI_ISL_623220\|20 | ···················· | ···················· | ····················· | ···················· | ···················· | ···················· |
| P | 1667 hCoV-19/England/MILK-ABB635/2020\|EPI_ISL_623221\|20 | ···················· | ···················· | ····················· | ···················· | ·····T·············· | ···················· |
| P | 1668 hCoV-19/England/MILK-AB7402/2020\|EPI_ISL_623223\|20 | ···················· | ···················· | ····················· | ···················· | ···················· | ···················· |
| P | 1669 hCoV-19/England/MILK-AB82E9/2020\|EPI_ISL_623225\|20 | ···················· | ···················· | ····················· | ···················· | ···················· | ···················· |
| P | 1670 hCoV-19/England/MILK-AB75B4/2020\|EPI_ISL_623226\|20 | ···················· | ···················· | ····················· | ···················· | ·····T·············· | ···················· |
| P | 1671 hCoV-19/England/MILK-AB8049/2020\|EPI_ISL_623227\|20 | ···················· | ···················· | ····················· | ···················· | ···················· | ···················· |
| P | 1672 hCoV-19/England/MILK-AB7675/2020\|EPI_ISL_623228\|20 | ···················· | ···················· | ····················· | ···················· | ···················· | ···················· |
| P | 1673 hCoV-19/England/ALDP-AA2464/2020\|EPI_ISL_624480\|20 | ···················· | ···················· | ····················· | ···················· | ···················· | ···················· |
| P | 1674 hCoV-19/England/ALDP-AA1A63/2020\|EPI_ISL_624481\|20 | ···················· | ·A·················· | ····················· | ···················· | ···················· | ··A················· |
| P | 1675 hCoV-19/England/ALDP-AA2868/2020\|EPI_ISL_624482\|20 | ···················· | ···················· | ····················· | ···················· | ···················· | ···················· |
| P | 1676 hCoV-19/England/ALDP-AA42FF/2020\|EPI_ISL_624483\|20 | ···················· | ···················· | ····················· | ···················· | ···················· | ···················· |
| P | 1677 hCoV-19/England/ALDP-AA1D5B/2020\|EPI_ISL_624485\|20 | ···················· | ···················· | ····················· | ···················· | ···················· | ···················· |
| P | 1678 hCoV-19/England/ALDP-AA215E/2020\|EPI_ISL_624486\|20 | ···················· | ···················· | ····················· | ···················· | ···················· | ···················· |
| P | 1679 hCoV-19/England/ALDP-AA209D/2020\|EPI_ISL_624487\|20 | ···················· | ···················· | ····················· | ···················· | ···················· | ···················· |
| P | 1680 hCoV-19/Wales/ALDP-AA4848/2020\|EPI_ISL_624489\|2020 | ···················· | ···················· | ····················· | ···················· | ···················· | ···················· |
| P | 1681 hCoV-19/England/ALDP-AA46A8/2020\|EPI_ISL_624490\|20 | ···················· | ·A·················· | ····················· | ···················· | ···················· | ··A················· |
| P | 1682 hCoV-19/England/ALDP-AA231C/2020\|EPI_ISL_624491\|20 | ···················· | ···················· | ····················· | ···················· | ···················· | ···················· |
| P | 1683 hCoV-19/Wales/ALDP-AA48A2/2020\|EPI_ISL_624492\|2020 | ···················· | ···················· | ····················· | ···················· | ···················· | ···················· |
| P | 1684 hCoV-19/England/ALDP-AA4BD6/2020\|EPI_ISL_624493\|20 | ···················· | ···················· | ····················· | ···················· | ···················· | ···················· |
| P | 1685 hCoV-19/England/ALDP-AA1F64/2020\|EPI_ISL_624494\|20 | ···················· | ···················· | ····················· | ···················· | ···················· | ···················· |
| P | 1686 hCoV-19/England/ALDP-AA233A/2020\|EPI_ISL_624495\|20 | ···················· | ···················· | ····················· | ···················· | ···················· | ···················· |
| P | 1687 hCoV-19/England/ALDP-AA15EA/2020\|EPI_ISL_624496\|20 | ···················· | ···················· | ····················· | ···················· | ···················· | ···················· |
| P | 1688 hCoV-19/England/ALDP-AA4286/2020\|EPI_ISL_624497\|20 | ···················· | ···················· | ····················· | ···················· | ···················· | ···················· |
| P | 1689 hCoV-19/England/ALDP-AA3C5C/2020\|EPI_ISL_624498\|20 | ···················· | ···················· | ····················· | ···················· | ···················· | ···················· |
| P | 1690 hCoV-19/England/ALDP-AA481B/2020\|EPI_ISL_624500\|20 | ···················· | ···················· | ····················· | ···················· | ···················· | ···················· |
| P | 1691 hCoV-19/England/ALDP-AA4611/2020\|EPI_ISL_624501\|20 | ···················· | ···················· | ····················· | ···················· | ···················· | ···················· |
| P | 1692 hCoV-19/England/ALDP-AA3DA4/2020\|EPI_ISL_624502\|20 | ···················· | ···················· | ····················· | ···················· | ···················· | ···················· |
| P | 1693 hCoV-19/England/ALDP-AA4417/2020\|EPI_ISL_624504\|20 | ···················· | ···················· | ····················· | ···················· | ···················· | ···················· |
| P | 1694 hCoV-19/England/ALDP-AA4347/2020\|EPI_ISL_624505\|20 | ···················· | ···················· | ····················· | ···················· | ···················· | ···················· |
| P | 1695 hCoV-19/England/ALDP-AA3E74/2020\|EPI_ISL_624506\|20 | ···················· | ···················· | ····················· | ···················· | ···················· | ···················· |
| P | 1696 hCoV-19/England/ALDP-AA1FEC/2020\|EPI_ISL_624507\|20 | ···················· | ···················· | ····················· | ···················· | ···················· | ···················· |
| P | 1697 hCoV-19/England/ALDP-AA1D97/2020\|EPI_ISL_624508\|20 | ···················· | ···················· | ····················· | ···················· | ···················· | ···················· |
| P | 1698 hCoV-19/Wales/ALDP-AA4884/2020\|EPI_ISL_624509\|2020 | ···················· | ···················· | ····················· | ···················· | ···················· | ···················· |
| P | 1699 hCoV-19/England/ALDP-AA3A16/2020\|EPI_ISL_624510\|20 | ···················· | ···················· | ····················· | ···················· | ···················· | ···················· |
| P | 1700 hCoV-19/England/ALDP-AA1A18/2020\|EPI_ISL_624511\|20 | ···················· | ···················· | ····················· | ···················· | ···················· | ···················· |
| P | 1701 hCoV-19/England/ALDP-AA1508/2020\|EPI_ISL_624512\|20 | ···················· | ···················· | ····················· | ···················· | ···················· | ···················· |
| P | 1702 hCoV-19/England/ALDP-AA159F/2020\|EPI_ISL_624513\|20 | ···················· | ···················· | ····················· | ···················· | ···················· | ···················· |
| P | 1703 hCoV-19/England/ALDP-AA4778/2020\|EPI_ISL_624515\|20 | ···················· | ···················· | ····················· | ···················· | ···················· | ···················· |
| P | 1704 hCoV-19/England/ALDP-AA1D4C/2020\|EPI_ISL_624516\|20 | ···················· | ···················· | ····················· | ···················· | ···················· | ···················· |
| P | 1705 hCoV-19/England/ALDP-AA3E29/2020\|EPI_ISL_624518\|20 | ···················· | ···················· | ····················· | ···················· | ···················· | ···················· |
| P | 1706 hCoV-19/England/ALDP-AA1025/2020\|EPI_ISL_624520\|20 | ···················· | ·A·················· | ····················· | ···················· | ···················· | ···················· |
| P | 1707 hCoV-19/England/ALDP-AA4699/2020\|EPI_ISL_624521\|20 | ···················· | ···················· | ····················· | ···················· | ···················· | ···················· |
| P | 1708 hCoV-19/England/ALDP-AA465D/2020\|EPI_ISL_624522\|20 | ···················· | ···················· | ····················· | ···················· | ···················· | ···················· |
| P | 1709 hCoV-19/England/ALDP-AA4839/2020\|EPI_ISL_624523\|20 | ···················· | ···················· | ····················· | ···················· | ···················· | ···················· |
| P | 1710 hCoV-19/England/ALDP-AA3C98/2020\|EPI_ISL_624524\|20 | ···················· | ···················· | ····················· | ···················· | ···················· | ···················· |
| P | 1711 hCoV-19/England/ALDP-AA19FD/2020\|EPI_ISL_624525\|20 | ···················· | ···················· | ····················· | ···················· | ···················· | ···················· |
| P | 1712 hCoV-19/England/ALDP-AA3AE9/2020\|EPI_ISL_624526\|20 | ···················· | ···················· | ····················· | ···················· | ···················· | ···················· |
| P | 1713 hCoV-19/England/ALDP-AA1AAF/2020\|EPI_ISL_624527\|20 | ···················· | ···················· | ····················· | ···················· | ···················· | ···················· |
| P | 1714 hCoV-19/England/ALDP-AA482A/2020\|EPI_ISL_624528\|20 | ···················· | ···················· | ····················· | ···················· | ···················· | ···················· |

| | | | | | | | | |
|---|---|---|---|---|---|---|---|---|
| P | 1715 hCoV-19/England/ALDP-AA4BC7/2020\|EPI_ISL_624529\|20 | ···················· | ···················· | ···················· | ···················· | ···················· | ···················· | |
| P | 1716 hCoV-19/England/ALDP-AA3C01/2020\|EPI_ISL_624531\|20 | ···················· | ···················· | ···················· | ···················· | ···················· | ···················· | |
| P | 1717 hCoV-19/England/ALDP-AA277A/2020\|EPI_ISL_624532\|20 | ···················· | ···················· | ···················· | ···················· | ···················· | ···················· | |
| P | 1718 hCoV-19/England/ALDP-AA3A25/2020\|EPI_ISL_624533\|20 | ···················· | ···················· | ···················· | ···················· | ···················· | ···················· | |
| P | 1719 hCoV-19/England/ALDP-AA3A8F/2020\|EPI_ISL_624534\|20 | ···················· | ···················· | ···················· | ···················· | ···················· | ···················· | |
| P | 1720 hCoV-19/England/ALDP-AA3E56/2020\|EPI_ISL_624535\|20 | ···················· | ···················· | ···················· | ···················· | ···················· | ···················· | |
| P | 1721 hCoV-19/Wales/ALDP-AA4866/2020\|EPI_ISL_624536\|2020 | ···················· | ···················· | ···················· | ···················· | ···················· | ···················· | |
| P | 1722 hCoV-19/England/ALDP-AA1CE5/2020\|EPI_ISL_624537\|20 | ···················· | ···················· | ···················· | ···················· | ···················· | ···················· | |
| P | 1723 hCoV-19/England/ALDP-AA3C4D/2020\|EPI_ISL_624539\|20 | ···················· | ···················· | ···················· | ···················· | ···················· | ···················· | |
| P | 1724 hCoV-19/England/ALDP-AA43ED/2020\|EPI_ISL_624540\|20 | ···················· | ···················· | ···················· | ···················· | ···················· | ···················· | |
| P | 1725 hCoV-19/England/ALDP-AA0F92/2020\|EPI_ISL_624541\|20 | ···················· | ···················· | ···················· | ···················· | ···················· | ···················· | |
| P | 1726 hCoV-19/England/ALDP-AA1104/2020\|EPI_ISL_624542\|20 | ···················· | ···················· | ···················· | ···················· | ···················· | ···················· | |
| P | 1727 hCoV-19/England/ALDP-AA42C2/2020\|EPI_ISL_624543\|20 | ···················· | ···················· | ···················· | ···················· | ···················· | ···················· | |
| P | 1728 hCoV-19/England/ALDP-AA48B1/2020\|EPI_ISL_624544\|20 | ···················· | ···················· | ···················· | ···················· | ···················· | ···················· | |
| P | 1729 hCoV-19/England/ALDP-AA2446/2020\|EPI_ISL_624545\|20 | ···················· | ···················· | ···················· | ···················· | ···················· | ···················· | |
| P | 1730 hCoV-19/England/ALDP-AA48EE/2020\|EPI_ISL_624547\|20 | ···················· | ···················· | ···················· | ···················· | ···················· | ···················· | |
| P | 1731 hCoV-19/England/ALDP-AA4BB8/2020\|EPI_ISL_624548\|20 | ···················· | ···················· | ···················· | ···················· | ···················· | ···················· | |
| P | 1732 hCoV-19/England/ALDP-AA39DD/2020\|EPI_ISL_624549\|20 | ···················· | ···················· | ···················· | ···················· | ···················· | ···················· | |
| P | 1733 hCoV-19/England/ALDP-AA3E83/2020\|EPI_ISL_624550\|20 | ···················· | ···················· | ···················· | ···················· | ···················· | ···················· | |
| P | 1734 hCoV-19/England/ALDP-AA1571/2020\|EPI_ISL_624551\|20 | ···················· | ···················· | ···················· | ···················· | ···················· | ···················· | |
| P | 1735 hCoV-19/England/ALDP-AA23FE/2020\|EPI_ISL_624552\|20 | ···················· | ···················· | ···················· | ···················· | ···················· | ···················· | |
| P | 1736 hCoV-19/England/ALDP-AA2033/2020\|EPI_ISL_624553\|20 | ···················· | ···················· | ···················· | ···················· | ···················· | ···················· | |
| P | 1737 hCoV-19/England/ALDP-AA46E4/2020\|EPI_ISL_624554\|20 | ···················· | ···················· | ···················· | ···················· | ···················· | ···················· | |
| P | 1738 hCoV-19/England/ALDP-AA0F74/2020\|EPI_ISL_624555\|20 | ···················· | ···················· | ···················· | ···················· | ···················· | ···················· | |
| P | 1739 hCoV-19/England/ALDP-AA3E1A/2020\|EPI_ISL_624556\|20 | ···················· | ···················· | ···················· | ···················· | ···················· | ···················· | |
| P | 1740 hCoV-19/England/ALDP-AA280E/2020\|EPI_ISL_624557\|20 | ···················· | ···················· | ···················· | ···················· | ···················· | ···················· | |
| P | 1741 hCoV-19/England/ALDP-AA4945/2020\|EPI_ISL_624559\|20 | ···················· | ···················· | ···················· | ···················· | ···················· | ···················· | |
| P | 1742 hCoV-19/Denmark/DCGC-8795/2020\|EPI_ISL_625803\|2020 | ···················· | ···················· | ···················· | ···················· | ···················· | ···················· | |
| P | 1743 hCoV-19/Denmark/DCGC-8745/2020\|EPI_ISL_625804\|2020 | ···················· | ···················· | ···················· | ···················· | ···················· | ···················· | |
| P | 1744 hCoV-19/Denmark/DCGC-8746/2020\|EPI_ISL_625805\|2020 | ···················· | ···················· | ···················· | ···················· | ···················· | ···················· | |
| P | 1745 hCoV-19/Denmark/DCGC-8772/2020\|EPI_ISL_625806\|2020 | ···················· | ···················· | ···················· | ···················· | ···················· | ···················· | |
| P | 1746 hCoV-19/Denmark/DCGC-8797/2020\|EPI_ISL_625807\|2020 | ···················· | ···················· | ···················· | ···················· | ···················· | ···················· | |
| P | 1747 hCoV-19/Denmark/DCGC-8750/2020\|EPI_ISL_625808\|2020 | ···················· | ···················· | ···················· | ···················· | ···················· | ···················· | |
| P | 1748 hCoV-19/Denmark/DCGC-8774/2020\|EPI_ISL_625809\|2020 | ···················· | ···················· | ···················· | ···················· | ···················· | ···················· | |
| P | 1749 hCoV-19/Denmark/DCGC-8786/2020\|EPI_ISL_625810\|2020 | ···················· | ···················· | ···················· | ···················· | ···················· | ···················· | |
| P | 1750 hCoV-19/Denmark/DCGC-8830/2020\|EPI_ISL_625811\|2020 | ···················· | ···················· | ···················· | ···················· | ···················· | ···················· | |
| P | 1751 hCoV-19/Denmark/DCGC-8865/2020\|EPI_ISL_625812\|2020 | ···················· | ···················· | ···················· | ···················· | ···················· | ···················· | |
| P | 1752 hCoV-19/Denmark/DCGC-8866/2020\|EPI_ISL_625813\|2020 | ···················· | ···················· | ···················· | ···················· | ···················· | ···················· | |
| P | 1753 hCoV-19/Denmark/DCGC-8828/2020\|EPI_ISL_625814\|2020 | ···················· | ···················· | ···················· | ···················· | ···················· | ···················· | |
| P | 1754 hCoV-19/Denmark/DCGC-8833/2020\|EPI_ISL_625815\|2020 | ···················· | ···················· | ···················· | ···················· | ···················· | ···················· | |
| P | 1755 hCoV-19/Denmark/DCGC-8843/2020\|EPI_ISL_625816\|2020 | ···················· | ···················· | ···················· | ···················· | ···················· | ···················· | |
| P | 1756 hCoV-19/Denmark/DCGC-8873/2020\|EPI_ISL_625817\|2020 | ···················· | ···················· | ···················· | ···················· | ···················· | ···················· | |
| P | 1757 hCoV-19/Denmark/DCGC-8888/2020\|EPI_ISL_625818\|2020 | ···················· | ···················· | ···················· | ···················· | ···················· | ···················· | |
| P | 1758 hCoV-19/Denmark/DCGC-8945/2020\|EPI_ISL_625819\|2020 | ···················· | ···················· | ···················· | ···················· | ···················· | ···················· | |
| P | 1759 hCoV-19/Denmark/DCGC-8881/2020\|EPI_ISL_625820\|2020 | ···················· | ···················· | ···················· | ···················· | ···················· | ···················· | |
| P | 1760 hCoV-19/Denmark/DCGC-8923/2020\|EPI_ISL_625821\|2020 | ···················· | ···················A | ···················· | ···················· | ···················· | ···················· | 1 |
| P | 1761 hCoV-19/Denmark/DCGC-8938/2020\|EPI_ISL_625822\|2020 | ···················· | ···················· | ···················· | ···················· | ···················· | ···················· | |
| P | 1762 hCoV-19/Denmark/DCGC-8896/2020\|EPI_ISL_625823\|2020 | ···················· | ···················· | ···················· | ···················· | ···················· | ···················· | |
| P | 1763 hCoV-19/Denmark/DCGC-8905/2020\|EPI_ISL_625824\|2020 | ···················· | ···················· | ···················· | ···················· | ···················· | ···················· | |
| P | 1764 hCoV-19/Denmark/DCGC-8926/2020\|EPI_ISL_625825\|2020 | ···················· | ···················· | ···················· | ···················· | ···················· | ···················· | |
| P | 1765 hCoV-19/Denmark/DCGC-8921/2020\|EPI_ISL_625826\|2020 | ···················· | ···················· | ···················· | ···················· | ···················· | ···················· | |
| P | 1766 hCoV-19/Denmark/DCGC-8956/2020\|EPI_ISL_625827\|2020 | ···················· | ···················· | ···················· | ···················· | ···················· | ···················· | |
| P | 1767 hCoV-19/Denmark/DCGC-8969/2020\|EPI_ISL_625828\|2020 | ···················· | ···················· | ···················· | ···················· | ···················· | ···················· | |
| P | 1768 hCoV-19/Denmark/DCGC-8985/2020\|EPI_ISL_625829\|2020 | ···················· | ···················· | ···················· | ···················· | ···················· | ···················· | |
| P | 1769 hCoV-19/Denmark/DCGC-8990/2020\|EPI_ISL_625830\|2020 | ···················· | ···················· | ···················· | ···················· | ···················· | ···················· | |
| P | 1770 hCoV-19/Denmark/DCGC-8999/2020\|EPI_ISL_625831\|2020 | ···················· | ···················· | ···················· | ···················· | ···················· | ···················· | |
| P | 1771 hCoV-19/Denmark/DCGC-8979/2020\|EPI_ISL_625832\|2020 | ···················· | ···················· | ···················· | ···················· | ···················· | ···················· | |
| P | 1772 hCoV-19/Denmark/DCGC-9009/2020\|EPI_ISL_625833\|2020 | ···················· | ···················· | ···················· | ···················· | ···················· | ···················· | |
| P | 1773 hCoV-19/Denmark/DCGC-8982/2020\|EPI_ISL_625834\|2020 | ···················· | ···················· | ···················· | ···················· | ···················· | ···················· | |
| P | 1774 hCoV-19/Denmark/DCGC-9011/2020\|EPI_ISL_625835\|2020 | ···················· | ···················· | ···················· | ···················· | ···················· | ···················· | |
| P | 1775 hCoV-19/Denmark/DCGC-8954/2020\|EPI_ISL_625836\|2020 | ···················· | ···················· | ···················· | ···················· | ···················· | ···················· | |
| P | 1776 hCoV-19/Denmark/DCGC-8975/2020\|EPI_ISL_625837\|2020 | ···················· | ···················· | ···················· | ···················· | ···················· | ···················· | |
| P | 1777 hCoV-19/Denmark/DCGC-8955/2020\|EPI_ISL_625838\|2020 | ···················· | ···················· | ···················· | ···················· | ···················· | ···················· | |
| P | 1778 hCoV-19/Denmark/DCGC-8997/2020\|EPI_ISL_625839\|2020 | ···················· | ···················· | ···················· | ···················· | ···················· | ···················· | |
| P | 1779 hCoV-19/Denmark/DCGC-8570/2020\|EPI_ISL_625842\|2020 | ···················· | ·A·················· | ···················· | ···················· | ···················· | ··A················· | |
| P | 1780 hCoV-19/Denmark/DCGC-8687/2020\|EPI_ISL_625844\|2020 | ···················· | ·A·················· | ···················· | ···················· | ···················· | ··A················· | |
| P | 1781 hCoV-19/Denmark/DCGC-8564/2020\|EPI_ISL_625846\|2020 | ···················· | ···················· | ···················· | ···················· | ···················· | ···················· | |
| P | 1782 hCoV-19/Denmark/DCGC-8552/2020\|EPI_ISL_625847\|2020 | ···················· | ···················· | ···················· | ···················· | ···················· | ···················· | |
| P | 1783 hCoV-19/Denmark/DCGC-8998/2020\|EPI_ISL_625848\|2020 | ···················· | ···················· | ···················· | ···················· | ···················· | ···················· | |
| P | 1784 hCoV-19/Denmark/DCGC-8650/2020\|EPI_ISL_625857\|2020 | ···················· | ···················· | ···················· | ···················· | ···················· | ···················· | |
| P | 1785 hCoV-19/Denmark/DCGC-8913/2020\|EPI_ISL_625872\|2020 | ···················· | ···················· | ···················· | ···················· | ···················· | ···················· | |
| P | 1786 hCoV-19/Denmark/DCGC-8588/2020\|EPI_ISL_625873\|2020 | ···················· | ···················· | ···················· | ···················· | ···················· | ···················· | |
| P | 1787 hCoV-19/Denmark/DCGC-8589/2020\|EPI_ISL_625874\|2020 | ···················· | ···················· | ···················· | ···················· | ···················· | ···················· | |
| P | 1788 hCoV-19/Denmark/DCGC-8604/2020\|EPI_ISL_625875\|2020 | ···················· | ·A·················· | ···················· | ···················· | ···················· | ··A················· | |
| P | 1789 hCoV-19/Denmark/DCGC-8657/2020\|EPI_ISL_625877\|2020 | ···················· | ·A·················· | ···················· | ···················· | ···················· | ··A················· | |
| P | 1790 hCoV-19/Denmark/DCGC-8658/2020\|EPI_ISL_625878\|2020 | ···················· | ···················· | ···················· | ···················· | ···················· | ···················· | |
| P | 1791 hCoV-19/Denmark/DCGC-8559/2020\|EPI_ISL_625885\|2020 | ···················· | ···················· | ···················· | ···················· | ···················· | ···················· | |
| P | 1792 hCoV-19/Denmark/DCGC-8659/2020\|EPI_ISL_625889\|2020 | ···················· | ···················· | ···················· | ···················· | ···················· | ···················· | |
| P | 1793 hCoV-19/Denmark/DCGC-3178/2020\|EPI_ISL_616000\|2020 | ···················· | ···················· | ···················· | ···················· | ···················· | ···················· | |
| P | 1794 hCoV-19/Denmark/DCGC-3179/2020\|EPI_ISL_616001\|2020 | ···················· | ···················· | ···················· | ···················· | ···················· | ···················· | |

| | | | | | | | | |
|---|---|---|---|---|---|---|---|---|
| P | 1795 | hCoV-19/Denmark/DCGC-3180/2020\|EPI_ISL_616002\|2020 | ···················· | ···················· | ···················· | ···················· | ···················· | ···················· |
| P | 1796 | hCoV-19/Denmark/DCGC-3181/2020\|EPI_ISL_616003\|2020 | ···················· | ···················· | ···················· | ···················· | ···················· | ···················· |
| P | 1797 | hCoV-19/Denmark/DCGC-3182/2020\|EPI_ISL_616004\|2020 | ···················· | ···················· | ···················· | ···················· | ···················· | ···················· |
| P | 1798 | hCoV-19/Denmark/DCGC-3183/2020\|EPI_ISL_616005\|2020 | ···················· | ···················· | ···················· | ···················· | ···················· | ···················· |
| P | 1799 | hCoV-19/Denmark/DCGC-3184/2020\|EPI_ISL_616006\|2020 | ···················· | ···················· | ···················· | ···················· | ···················· | ···················· |
| P | 1800 | hCoV-19/Denmark/DCGC-3185/2020\|EPI_ISL_616007\|2020 | ···················· | ···················· | ···················· | ···················· | ···················· | ···················· |
| P | 1801 | hCoV-19/Denmark/DCGC-3186/2020\|EPI_ISL_616008\|2020 | ···················· | ···················· | ···················· | ···················· | ···················· | ···················· |
| P | 1802 | hCoV-19/Denmark/DCGC-3187/2020\|EPI_ISL_616009\|2020 | ···················· | ···················· | ···················· | ···················· | ···················· | ···················· |
| P | 1803 | hCoV-19/Denmark/DCGC-3253/2020\|EPI_ISL_616010\|2020 | ···················· | ···················· | ···················· | ···················· | ···················· | ···················· |
| P | 1804 | hCoV-19/Denmark/DCGC-3190/2020\|EPI_ISL_616013\|2020 | ···················· | ···················· | ···················· | ···················· | ···················· | ···················· |
| P | 1805 | hCoV-19/Denmark/DCGC-3191/2020\|EPI_ISL_616014\|2020 | ···················· | ···················· | ···················· | ···················· | ···················· | ···················· |
| P | 1806 | hCoV-19/Denmark/DCGC-3192/2020\|EPI_ISL_616015\|2020 | ···················· | ···················· | ···················· | ···················· | ···················· | ···················· |
| P | 1807 | hCoV-19/Denmark/DCGC-3193/2020\|EPI_ISL_616016\|2020 | ···················· | ···················· | ···················· | ···················· | ···················· | ···················· |
| P | 1808 | hCoV-19/Denmark/DCGC-3194/2020\|EPI_ISL_616017\|2020 | ···················· | ···················· | ···················· | ···················· | ···················· | ···················· |
| P | 1809 | hCoV-19/Denmark/DCGC-3195/2020\|EPI_ISL_616018\|2020 | ···················· | ···················· | ···················· | ···················· | ···················· | ···················· |
| P | 1810 | hCoV-19/Denmark/DCGC-3196/2020\|EPI_ISL_616019\|2020 | ···················· | ···················· | ···················· | ···················· | ···················· | ···················· |
| P | 1811 | hCoV-19/Denmark/DCGC-8294/2020\|EPI_ISL_617310\|2020 | ···················· | ···················· | ···················· | ···················· | ···················· | ···················· |
| P | 1812 | hCoV-19/Denmark/DCGC-8308/2020\|EPI_ISL_617311\|2020 | ···················· | ···················· | ···················· | ···················· | ···················· | ···················· |
| P | 1813 | hCoV-19/Denmark/DCGC-8326/2020\|EPI_ISL_617312\|2020 | ···················· | ···················· | ···················· | ···················· | ···················· | ···················· |
| P | 1814 | hCoV-19/Denmark/DCGC-8333/2020\|EPI_ISL_617313\|2020 | ···················· | ···················· | ···················· | ···················· | ···················· | ···················· |
| P | 1815 | hCoV-19/Denmark/DCGC-8263/2020\|EPI_ISL_617314\|2020 | ···················· | ···················· | ···················· | ···················· | ···················· | ···················· |
| P | 1816 | hCoV-19/Denmark/DCGC-8287/2020\|EPI_ISL_617315\|2020 | ···················· | ···················· | ···················· | ···················· | ···················· | ···················· |
| P | 1817 | hCoV-19/Denmark/DCGC-8303/2020\|EPI_ISL_617316\|2020 | ···················· | ···················· | ···················· | ···················· | ···················· | ···················· |
| P | 1818 | hCoV-19/Denmark/DCGC-8320/2020\|EPI_ISL_617317\|2020 | ···················· | ···················· | ···················· | ···················· | ···················· | ···················· |
| P | 1819 | hCoV-19/Denmark/DCGC-8264/2020\|EPI_ISL_617318\|2020 | ···················· | ···················· | ···················· | ···················· | ···················· | ···················· |
| P | 1820 | hCoV-19/Denmark/DCGC-8272/2020\|EPI_ISL_617319\|2020 | ···················· | ···················· | ···················· | ···················· | ···················· | ···················· |
| P | 1821 | hCoV-19/Denmark/DCGC-8283/2020\|EPI_ISL_617320\|2020 | ···················· | ···················· | ···················· | ···················· | ···················· | ···················· |
| P | 1822 | hCoV-19/Denmark/DCGC-8296/2020\|EPI_ISL_617321\|2020 | ···················· | ···················· | ···················· | ···················· | ···················· | ···················· |
| P | 1823 | hCoV-19/Denmark/DCGC-8304/2020\|EPI_ISL_617322\|2020 | ···················· | ···················· | ···················· | ···················· | ···················· | ···················· |
| P | 1824 | hCoV-19/Denmark/DCGC-8309/2020\|EPI_ISL_617323\|2020 | ···················· | ···················· | ···················· | ···················· | ···················· | ···················· |
| P | 1825 | hCoV-19/Denmark/DCGC-8265/2020\|EPI_ISL_617324\|2020 | ···················· | ···················· | ···················· | ···················· | ···················· | ···················· |
| P | 1826 | hCoV-19/Denmark/DCGC-8279/2020\|EPI_ISL_617325\|2020 | ···················· | ···················· | ···················· | ···················· | ···················· | ···················· |
| P | 1827 | hCoV-19/Denmark/DCGC-8284/2020\|EPI_ISL_617326\|2020 | ···················· | ···················· | ···················· | ···················· | ···················· | ···················· |
| P | 1828 | hCoV-19/Denmark/DCGC-8314/2020\|EPI_ISL_617327\|2020 | ···················· | ···················· | ···················· | ···················· | ···················· | ···················· |
| P | 1829 | hCoV-19/Denmark/DCGC-8322/2020\|EPI_ISL_617328\|2020 | ···················· | ···················· | ···················· | ···················· | ···················· | ···················· |
| P | 1830 | hCoV-19/Denmark/DCGC-8328/2020\|EPI_ISL_617329\|2020 | ···················· | ···················· | ···················· | ···················· | ···················· | ···················· |
| P | 1831 | hCoV-19/Denmark/DCGC-8340/2020\|EPI_ISL_617330\|2020 | ···················· | ···················· | ···················· | ···················· | ···················· | ···················· |
| P | 1832 | hCoV-19/Denmark/DCGC-8346/2020\|EPI_ISL_617331\|2020 | ···················· | ···················· | ···················· | ···················· | ···················· | ···················· |
| P | 1833 | hCoV-19/Denmark/DCGC-8393/2020\|EPI_ISL_617332\|2020 | ···················· | ···················· | ···················· | ···················· | ···················· | ···················· |
| P | 1834 | hCoV-19/Denmark/DCGC-8400/2020\|EPI_ISL_617333\|2020 | ···················· | ···················· | ···················· | ···················· | ···················· | ···················· |
| P | 1835 | hCoV-19/Denmark/DCGC-8334/2020\|EPI_ISL_617334\|2020 | ···················· | ···················· | ···················· | ···················· | ···················· | ···················· |
| P | 1836 | hCoV-19/Denmark/DCGC-8341/2020\|EPI_ISL_617335\|2020 | ···················· | ···················· | ···················· | ···················· | ···················· | ···················· |
| P | 1837 | hCoV-19/Denmark/DCGC-8347/2020\|EPI_ISL_617336\|2020 | ···················· | ···················· | ···················· | ···················· | ···················· | ···················· |
| P | 1838 | hCoV-19/Denmark/DCGC-8354/2020\|EPI_ISL_617337\|2020 | ···················· | ···················· | ···················· | ···················· | ···················· | ···················· |
| P | 1839 | hCoV-19/Denmark/DCGC-8369/2020\|EPI_ISL_617338\|2020 | ···················· | ···················· | ···················· | ···················· | ···················· | ···················· |
| P | 1840 | hCoV-19/Denmark/DCGC-8376/2020\|EPI_ISL_617339\|2020 | ···················· | ···················· | ···················· | ···················· | ···················· | ···················· |
| P | 1841 | hCoV-19/Denmark/DCGC-8381/2020\|EPI_ISL_617340\|2020 | ···················· | ·A·················· | ···················· | ···················· | ···················· | ···················· |
| P | 1842 | hCoV-19/Denmark/DCGC-8386/2020\|EPI_ISL_617341\|2020 | ···················· | ···················· | ···················· | ···················· | ···················· | ···················· |
| P | 1843 | hCoV-19/Denmark/DCGC-8335/2020\|EPI_ISL_617343\|2020 | ···················· | ···················· | ···················· | ···················· | ···················· | ···················· |
| P | 1844 | hCoV-19/Denmark/DCGC-8342/2020\|EPI_ISL_617344\|2020 | ···················· | ···················· | ···················· | ···················· | ···················· | ···················· |
| P | 1845 | hCoV-19/Denmark/DCGC-8366/2020\|EPI_ISL_617345\|2020 | ···················· | ···················· | ···················· | ···················· | ···················· | ···················· |
| P | 1846 | hCoV-19/Denmark/DCGC-8370/2020\|EPI_ISL_617346\|2020 | ···················· | ···················· | ···················· | ···················· | ···················· | ···················· |
| P | 1847 | hCoV-19/Denmark/DCGC-8377/2020\|EPI_ISL_617347\|2020 | ···················· | ···················· | ···················· | ···················· | ···················· | ···················· |
| P | 1848 | hCoV-19/Denmark/DCGC-8387/2020\|EPI_ISL_617348\|2020 | ···················· | ···················· | ···················· | ···················· | ···················· | ···················· |
| P | 1849 | hCoV-19/Denmark/DCGC-8336/2020\|EPI_ISL_617349\|2020 | ···················· | ···················· | ···················· | ···················· | ···················· | ···················· |
| P | 1850 | hCoV-19/Denmark/DCGC-1596/2020\|EPI_ISL_618640\|2020 | ···················· | ···················· | ···················· | ···················· | ···················· | ···················· |
| P | 1851 | hCoV-19/Denmark/DCGC-1560/2020\|EPI_ISL_618641\|2020 | ···················· | ···················· | ···················· | ···················· | ···················· | ···················· |
| P | 1852 | hCoV-19/Denmark/DCGC-1561/2020\|EPI_ISL_618642\|2020 | ···················· | ···················· | ···················· | ···················· | ···················· | ···················· |
| P | 1853 | hCoV-19/Denmark/DCGC-1597/2020\|EPI_ISL_618643\|2020 | ···················· | ···················· | ···················· | ···················· | ···················· | ···················· |
| P | 1854 | hCoV-19/Denmark/DCGC-1598/2020\|EPI_ISL_618644\|2020 | ···················· | ···················· | ···················· | ···················· | ···················· | ···················· |
| P | 1855 | hCoV-19/Denmark/DCGC-1599/2020\|EPI_ISL_618645\|2020 | ···················· | ···················· | ···················· | ···················· | ···················· | ···················· |
| P | 1856 | hCoV-19/Denmark/DCGC-1601/2020\|EPI_ISL_618647\|2020 | ···················· | ···················· | ···················· | ···················· | ···················· | ···················· |
| P | 1857 | hCoV-19/Denmark/DCGC-1602/2020\|EPI_ISL_618648\|2020 | ···················· | ···················· | ···················· | ···················· | ···················· | ···················· |
| P | 1858 | hCoV-19/Denmark/DCGC-1603/2020\|EPI_ISL_618649\|2020 | ···················· | ···················· | ···················· | ···················· | ···················· | ···················· |
| P | 1859 | hCoV-19/Denmark/DCGC-1604/2020\|EPI_ISL_618650\|2020 | ···················· | ···················· | ···················· | ···················· | ···················· | ···················· |
| P | 1860 | hCoV-19/Denmark/DCGC-1605/2020\|EPI_ISL_618651\|2020 | ···················· | ···················· | ···················· | ···················· | ···················· | ···················· |
| P | 1861 | hCoV-19/Denmark/DCGC-1562/2020\|EPI_ISL_618652\|2020 | ···················· | ···················· | ···················· | ···················· | ···················· | ···················· |
| P | 1862 | hCoV-19/Denmark/DCGC-1548/2020\|EPI_ISL_618653\|2020 | ···················· | ···················· | ···················· | ···················· | ···················· | ···················· |
| P | 1863 | hCoV-19/Denmark/DCGC-1549/2020\|EPI_ISL_618654\|2020 | ···················· | ···················· | ···················· | ···················· | ···················· | ···················· |
| P | 1864 | hCoV-19/Denmark/DCGC-1550/2020\|EPI_ISL_618655\|2020 | ···················· | ···················· | ···················· | ···················· | ···················· | ···················· |
| P | 1865 | hCoV-19/Denmark/DCGC-1551/2020\|EPI_ISL_618656\|2020 | ···················· | ···················· | ···················· | ···················· | ···················· | ···················· |
| P | 1866 | hCoV-19/Denmark/DCGC-1552/2020\|EPI_ISL_618657\|2020 | ···················· | ···················· | ···················· | ···················· | ···················· | ···················· |
| P | 1867 | hCoV-19/Denmark/DCGC-1554/2020\|EPI_ISL_618659\|2020 | ···················· | ···················· | ···················· | ···················· | ···················· | ···················· |
| P | 1868 | hCoV-19/Denmark/DCGC-1556/2020\|EPI_ISL_618661\|2020 | ···················· | ···················· | ···················· | ···················· | ···················· | ···················· |
| P | 1869 | hCoV-19/Denmark/DCGC-1388/2020\|EPI_ISL_618662\|2020 | ···················· | ···················· | ···················· | ···················· | ···················· | ···················· |
| P | 1870 | hCoV-19/Denmark/DCGC-7721/2020\|EPI_ISL_618663\|2020 | ···················· | ···················· | ···················· | ···················· | ···················· | ···················· |
| P | 1871 | hCoV-19/Denmark/DCGC-7729/2020\|EPI_ISL_618664\|2020 | ···················· | ···················· | ···················· | ···················· | ···················· | ···················· |
| P | 1872 | hCoV-19/Denmark/DCGC-7733/2020\|EPI_ISL_618665\|2020 | ···················· | ···················· | ···················· | ···················· | ···················· | ···················· |
| P | 1873 | hCoV-19/Denmark/DCGC-7741/2020\|EPI_ISL_618666\|2020 | ···················· | ·A·················· | ···················· | ···················· | ···················· | ···A················ |
| P | 1874 | hCoV-19/Denmark/DCGC-7751/2020\|EPI_ISL_618668\|2020 | ···················· | ···················· | ···················· | ···················· | ···················· | ···················· |

| | | | | | | | |
|---|---|---|---|---|---|---|---|
| P | 1875 hCoV-19/Denmark/DCGC-7758/2020\|EPI_ISL_618669\|2020 | ···················· | ···················· | ····················· | ···················· | ···················· | ···················· |
| P | 1876 hCoV-19/Denmark/DCGC-7776/2020\|EPI_ISL_618671\|2020 | ···················· | ···················· | ····················· | ···················· | ···················· | ···················· |
| P | 1877 hCoV-19/Denmark/DCGC-7730/2020\|EPI_ISL_618675\|2020 | ···················· | ···················· | ····················· | ···················· | ···················· | ···················· |
| P | 1878 hCoV-19/Denmark/DCGC-7734/2020\|EPI_ISL_618676\|2020 | ···················· | ···················· | ····················· | ···················· | ···················· | ···················· |
| P | 1879 hCoV-19/Denmark/DCGC-7742/2020\|EPI_ISL_618677\|2020 | ···················· | ·A·················· | ····················· | ···················· | ···················· | ··A················· |
| P | 1880 hCoV-19/Denmark/DCGC-7752/2020\|EPI_ISL_618678\|2020 | ···················· | ·A·················· | ····················· | ···················· | ···················· | ···················· |
| P | 1881 hCoV-19/Denmark/DCGC-7759/2020\|EPI_ISL_618679\|2020 | ···················· | ···················· | ····················· | ···················· | ···················· | ···················· |
| P | 1882 hCoV-19/Denmark/DCGC-5631/2020\|EPI_ISL_619918\|2020 | ···················· | ···················· | ····················· | ···················· | ···················· | ···················· |
| P | 1883 hCoV-19/Denmark/DCGC-5632/2020\|EPI_ISL_619919\|2020 | ···················· | ···················· | ····················· | ···················· | ···················· | ···················· |
| P | 1884 hCoV-19/Denmark/DCGC-5633/2020\|EPI_ISL_619920\|2020 | ···················· | ···················· | ····················· | ···················· | ···················· | ···················· |
| P | 1885 hCoV-19/Denmark/DCGC-5634/2020\|EPI_ISL_619921\|2020 | ···················· | ···················· | ····················· | ···················· | ···················· | ···················· |
| P | 1886 hCoV-19/Denmark/DCGC-5635/2020\|EPI_ISL_619922\|2020 | ···················· | ···················· | ····················· | ···················· | ···················· | ···················· |
| P | 1887 hCoV-19/Denmark/DCGC-5636/2020\|EPI_ISL_619923\|2020 | ···················· | ···················· | ····················· | ···················· | ···················· | ···················· |
| P | 1888 hCoV-19/Denmark/DCGC-5637/2020\|EPI_ISL_619924\|2020 | ···················· | ···················· | ····················· | ···················· | ···················· | ···················· |
| P | 1889 hCoV-19/Denmark/DCGC-5638/2020\|EPI_ISL_619925\|2020 | ···················· | ···················· | ····················· | ···················· | ···················· | ···················· |
| P | 1890 hCoV-19/Denmark/DCGC-5640/2020\|EPI_ISL_619927\|2020 | ···················· | ···················· | ····················· | ···················· | ···················· | ···················· |
| P | 1891 hCoV-19/Denmark/DCGC-5641/2020\|EPI_ISL_619928\|2020 | ···················· | ···················· | ····················· | ···················· | ···················· | ···················· |
| P | 1892 hCoV-19/Denmark/DCGC-5683/2020\|EPI_ISL_619970\|2020 | ···················· | ···················· | ····················· | ···················· | ···················· | ···················· |
| P | 1893 hCoV-19/Denmark/DCGC-5684/2020\|EPI_ISL_619971\|2020 | ···················· | ···················· | ····················· | ···················· | ···················· | ···················· |
| P | 1894 hCoV-19/Denmark/DCGC-5685/2020\|EPI_ISL_619972\|2020 | ···················· | ·A·················· | ····················· | ···················· | ···················· | ···················· |
| P | 1895 hCoV-19/Denmark/DCGC-5686/2020\|EPI_ISL_619973\|2020 | ···················· | ···················· | ····················· | ···················· | ···················· | ···················· |
| P | 1896 hCoV-19/Denmark/DCGC-5688/2020\|EPI_ISL_619975\|2020 | ···················· | ···················· | ····················· | ···················· | ···················· | ···················· |
| P | 1897 hCoV-19/Denmark/DCGC-5689/2020\|EPI_ISL_619976\|2020 | ···················· | ···················· | ····················· | ···················· | ···················· | ···················· |
| P | 1898 hCoV-19/Denmark/DCGC-1746/2020\|EPI_ISL_619978\|2020 | ···················· | ···················· | ····················· | ···················· | ···················· | ···················· |
| P | 1899 hCoV-19/Denmark/DCGC-1734/2020\|EPI_ISL_619979\|2020 | ···················· | ···················· | ····················· | ···················· | ···················· | ···················· |
| P | 1900 hCoV-19/Denmark/DCGC-1739/2020\|EPI_ISL_619980\|2020 | ···················· | ···················· | ····················· | ···················· | ···················· | ···················· |
| P | 1901 hCoV-19/Denmark/DCGC-1740/2020\|EPI_ISL_619981\|2020 | ···················· | ···················· | ····················· | ···················· | ···················· | ···················· |
| P | 1902 hCoV-19/Denmark/DCGC-1742/2020\|EPI_ISL_619982\|2020 | ···················· | ···················· | ····················· | ···················· | ···················· | ···················· |
| P | 1903 hCoV-19/Denmark/DCGC-1745/2020\|EPI_ISL_619983\|2020 | ···················· | ···················· | ····················· | ···················· | ···················· | ···················· |
| P | 1904 hCoV-19/Denmark/DCGC-6226/2020\|EPI_ISL_619986\|2020 | ···················· | ···················· | ····················· | ···················· | ···················· | ···················· |
| P | 1905 hCoV-19/Denmark/DCGC-6229/2020\|EPI_ISL_619989\|2020 | ···················· | ···················· | ····················· | ···················· | ···················· | ···················· |
| P | 1906 hCoV-19/Denmark/DCGC-6231/2020\|EPI_ISL_619991\|2020 | ···················· | ···················· | ····················· | ···················· | ···················· | ···················· |
| P | 1907 hCoV-19/Denmark/DCGC-6232/2020\|EPI_ISL_619992\|2020 | ···················· | ···················· | ····················· | ···················· | ···················· | ···················· |
| P | 1908 hCoV-19/Denmark/DCGC-6233/2020\|EPI_ISL_619993\|2020 | ···················· | ···················· | ····················· | ···················· | ···················· | ···················· |
| P | 1909 hCoV-19/Denmark/DCGC-6234/2020\|EPI_ISL_619994\|2020 | ···················· | ···················· | ····················· | ···················· | ···················· | ···················· |
| P | 1910 hCoV-19/Denmark/DCGC-6235/2020\|EPI_ISL_619995\|2020 | ···················· | ···················· | ····················· | ···················· | ···················· | ···················· |
| P | 1911 hCoV-19/Denmark/DCGC-6236/2020\|EPI_ISL_619996\|2020 | ···················· | ···················· | ····················· | ···················· | ···················· | ···················· |
| P | 1912 hCoV-19/Denmark/DCGC-6237/2020\|EPI_ISL_619997\|2020 | ···················· | ···················· | ····················· | ···················· | ···················· | ···················· |
| P | 1913 hCoV-19/Denmark/DCGC-6239/2020\|EPI_ISL_619999\|2020 | ···················· | ···················· | ····················· | ···················· | ···················· | ···················· |
| P | 1914 hCoV-19/Denmark/DCGC-5765/2020\|EPI_ISL_620952\|2020 | ···················· | ···················· | ····················· | ···················· | ···················· | ···················· |
| P | 1915 hCoV-19/Denmark/DCGC-5767/2020\|EPI_ISL_620954\|2020 | ···················· | ···················· | ····················· | ···················· | ···················· | ···················· |
| P | 1916 hCoV-19/Denmark/DCGC-5768/2020\|EPI_ISL_620955\|2020 | ···················· | ···················· | ····················· | ···················· | ···················· | ···················· |
| P | 1917 hCoV-19/Denmark/DCGC-5769/2020\|EPI_ISL_620956\|2020 | ···················· | ···················· | ····················· | ···················· | ···················· | ···················· |
| P | 1918 hCoV-19/Denmark/DCGC-5770/2020\|EPI_ISL_620957\|2020 | ···················· | ···················· | ····················· | ···················· | ···················· | ···················· |
| P | 1919 hCoV-19/Denmark/DCGC-5772/2020\|EPI_ISL_620959\|2020 | ···················· | ···················· | ····················· | ···················· | ···················· | ···················· |
| P | 1920 hCoV-19/Denmark/DCGC-5773/2020\|EPI_ISL_620960\|2020 | ···················· | ···················· | ····················· | ···················· | ···················· | ···················· |
| P | 1921 hCoV-19/Denmark/DCGC-5774/2020\|EPI_ISL_620961\|2020 | ···················· | ···················· | ····················· | ···················· | ···················· | ···················· |
| P | 1922 hCoV-19/Denmark/DCGC-5775/2020\|EPI_ISL_620962\|2020 | ···················· | ···················· | ····················· | ···················· | ···················· | ···················· |
| P | 1923 hCoV-19/Denmark/DCGC-5776/2020\|EPI_ISL_620963\|2020 | ···················· | ···················· | ····················· | ···················· | ···················· | ···················· |
| P | 1924 hCoV-19/Denmark/DCGC-5777/2020\|EPI_ISL_620964\|2020 | ···················· | ···················· | ····················· | ···················· | ···················· | ···················· |
| P | 1925 hCoV-19/Denmark/DCGC-5778/2020\|EPI_ISL_620965\|2020 | ···················· | ···················· | ····················· | ···················· | ···················· | ···················· |
| P | 1926 hCoV-19/Denmark/DCGC-5779/2020\|EPI_ISL_620966\|2020 | ···················· | ···················· | ····················· | ···················· | ···················· | ···················· |
| P | 1927 hCoV-19/Denmark/DCGC-5780/2020\|EPI_ISL_620967\|2020 | ···················· | ···················· | ····················· | ···················· | ···················· | ···················· |
| P | 1928 hCoV-19/Denmark/DCGC-5781/2020\|EPI_ISL_620968\|2020 | ···················· | ···················· | ····················· | ···················· | ···················· | ···················· |
| P | 1929 hCoV-19/Denmark/DCGC-5782/2020\|EPI_ISL_620969\|2020 | ···················· | ···················· | ····················· | ···················· | ···················· | ···················· |
| P | 1930 hCoV-19/Denmark/DCGC-5783/2020\|EPI_ISL_620970\|2020 | ···················· | ···················· | ····················· | ···················· | ···················· | ···················· |
| P | 1931 hCoV-19/Denmark/DCGC-5784/2020\|EPI_ISL_620971\|2020 | ···················· | ···················· | ····················· | ···················· | ···················· | ···················· |
| P | 1932 hCoV-19/Denmark/DCGC-5786/2020\|EPI_ISL_620973\|2020 | ···················· | ···················· | ····················· | ···················· | ···················· | ···················· |
| P | 1933 hCoV-19/Denmark/DCGC-5787/2020\|EPI_ISL_620974\|2020 | ···················· | ···················· | ····················· | ···················· | ···················· | ···················· |
| P | 1934 hCoV-19/Denmark/DCGC-5789/2020\|EPI_ISL_620976\|2020 | ···················· | ···················· | ····················· | ···················· | ···················· | ···················· |
| P | 1935 hCoV-19/Denmark/DCGC-5790/2020\|EPI_ISL_620977\|2020 | ···················· | ···················· | ····················· | ···················· | ···················· | ···················· |
| P | 1936 hCoV-19/Denmark/DCGC-5791/2020\|EPI_ISL_620978\|2020 | ···················· | ···················· | ····················· | ···················· | ···················· | ···················· |
| P | 1937 hCoV-19/Denmark/DCGC-5792/2020\|EPI_ISL_620979\|2020 | ···················· | ···················· | ····················· | ···················· | ···················· | ···················· |
| P | 1938 hCoV-19/Denmark/DCGC-5798/2020\|EPI_ISL_620985\|2020 | ···················· | ···················· | ····················· | ···················· | ···················· | ···················· |
| P | 1939 hCoV-19/Denmark/DCGC-5799/2020\|EPI_ISL_620986\|2020 | ···················· | ···················· | ····················· | ···················· | ···················· | ···················· |
| P | 1940 hCoV-19/Denmark/DCGC-5800/2020\|EPI_ISL_620987\|2020 | ···················· | ···················· | ····················· | ···················· | ···················· | ···················· |
| P | 1941 hCoV-19/Denmark/DCGC-5801/2020\|EPI_ISL_620988\|2020 | ···················· | ···················· | ····················· | ···················· | ···················· | ···················· |
| P | 1942 hCoV-19/Denmark/DCGC-5802/2020\|EPI_ISL_620989\|2020 | ···················· | ···················· | ····················· | ···················· | ···················· | ···················· |
| P | 1943 hCoV-19/Denmark/DCGC-5803/2020\|EPI_ISL_620990\|2020 | ···················· | ···················· | ····················· | ···················· | ···················· | ···················· |
| P | 1944 hCoV-19/Denmark/DCGC-5804/2020\|EPI_ISL_620991\|2020 | ···················· | ···················· | ····················· | ···················· | ···················· | ···················· |
| P | 1945 hCoV-19/Denmark/DCGC-5805/2020\|EPI_ISL_620992\|2020 | ···················· | ···················· | ····················· | ···················· | ···················· | ···················· |
| P | 1946 hCoV-19/Denmark/DCGC-5806/2020\|EPI_ISL_620993\|2020 | ···················· | ···················· | ····················· | ···················· | ···················· | ···················· |
| P | 1947 hCoV-19/Denmark/DCGC-5807/2020\|EPI_ISL_620994\|2020 | ···················· | ···················· | ····················· | ···················· | ···················· | ···················· |
| P | 1948 hCoV-19/Denmark/DCGC-5808/2020\|EPI_ISL_620995\|2020 | ···················· | ···················· | ····················· | ···················· | ···················· | ···················· |
| P | 1949 hCoV-19/Brazil/RJ-00305/2020\|EPI_ISL_623108\|2020-0 | ···················· | ···················· | ····················· | ···················· | ···················· | ···················· |
| P | 1950 hCoV-19/Brazil/RJ-00307/2020\|EPI_ISL_623110\|2020-0 | ···················· | ···················· | ····················· | ···················· | ···················· | ···················· |
| P | 1951 hCoV-19/Brazil/RJ-00309/2020\|EPI_ISL_623112\|2020-0 | ···················· | ···················· | ····················· | ···················· | ···················· | ···················· |
| P | 1952 hCoV-19/Brazil/RJ-00311/2020\|EPI_ISL_623114\|2020-0 | ···················· | ···················· | ····················· | ···················· | ···················· | ···················· |
| P | 1953 hCoV-19/Brazil/RJ-00313/2020\|EPI_ISL_623116\|2020-0 | ···················· | ···················· | ····················· | ···················· | ···················· | ···················· |
| P | 1954 hCoV-19/Brazil/RJ-00315/2020\|EPI_ISL_623118\|2020-0 | ···················· | ···················· | ····················· | ···················· | ···················· | ···················· |

| | | | | | | | |
|---|---|---|---|---|---|---|---|
| P | 1955 hCoV-19/Brazil/RJ-00316/2020\|EPI_ISL_623119\|2020-0 | ···················· | ···················· | ····················· | ···················· | ···················· | ···················· |
| P | 1956 hCoV-19/Brazil/RJ-00317/2020\|EPI_ISL_623120\|2020-0 | ···················· | ···················· | ····················· | ···················· | ···················· | ···················· |
| P | 1957 hCoV-19/Brazil/RJ-00321/2020\|EPI_ISL_623124\|2020-0 | ···················· | ···················· | ····················· | ···················· | ···················· | ···················· |
| P | 1958 hCoV-19/Brazil/RJ-00323/2020\|EPI_ISL_623126\|2020-0 | ···················· | ···················· | ····················· | ···················· | ···················· | ···················· |
| P | 1960 hCoV-19/Brazil/RJ-00327/2020\|EPI_ISL_623130\|2020-0 | ···················· | ···················· | ····················· | ···················· | ···················· | ···················· |
| P | 1961 hCoV-19/Brazil/RJ-00329/2020\|EPI_ISL_623132\|2020-0 | ···················· | ···················· | ····················· | ···················· | ···················· | ···················· |
| P | 1962 hCoV-19/Brazil/RJ-00331/2020\|EPI_ISL_623134\|2020-0 | ···················· | ···················· | ····················· | ···················· | ···················· | ···················· |
| P | 1963 hCoV-19/Brazil/RJ-00333/2020\|EPI_ISL_623136\|2020-0 | ···················· | ···················· | ····················· | ···················· | ···················· | ···················· |
| P | 1964 hCoV-19/Brazil/RJ-00335/2020\|EPI_ISL_623138\|2020-0 | ···················· | ···················· | ····················· | ···················· | ···················· | ···················· |
| P | 1965 hCoV-19/Brazil/RJ-00337/2020\|EPI_ISL_623140\|2020-0 | ···················· | ···················· | ····················· | ···················· | ···················· | ···················· |
| P | 1966 hCoV-19/Brazil/RJ-00339/2020\|EPI_ISL_623142\|2020-0 | ···················· | ···················· | ····················· | ···················· | ···················· | ···················· |
| P | 1967 hCoV-19/Brazil/RJ-00340/2020\|EPI_ISL_623143\|2020-0 | ···················· | ···················· | ····················· | ···················· | ···················· | ···················· |
| P | 1968 hCoV-19/Brazil/RJ-00341/2020\|EPI_ISL_623144\|2020-0 | ···················· | ···················· | ····················· | ···················· | ···················· | ···················· |
| P | 1969 hCoV-19/Brazil/RJ-00342/2020\|EPI_ISL_623145\|2020-0 | ···················· | ···················· | ····················· | ···················· | ···················· | ···················· |
| P | 1970 hCoV-19/Brazil/RJ-00343/2020\|EPI_ISL_623146\|2020-0 | ···················· | ···················· | ····················· | ···················· | ···················· | ···················· |
| P | 1971 hCoV-19/Brazil/RJ-00344/2020\|EPI_ISL_623147\|2020-0 | ···················· | ···················· | ····················· | ···················· | ···················· | ···················· |
| P | 1972 hCoV-19/Brazil/RJ-00345/2020\|EPI_ISL_623148\|2020-0 | ···················· | ···················· | ····················· | ···················· | ···················· | ···················· |
| P | 1973 hCoV-19/Brazil/RJ-00346/2020\|EPI_ISL_623149\|2020-0 | ···················· | ···················· | ····················· | ···················· | ···················· | ···················· |
| P | 1974 hCoV-19/USA/UT-UPHL-201103917/2020\|EPI_ISL_623171\| | ···················· | ···················· | ····················· | ···················· | ···················· | ···················· |
| P | 1975 hCoV-19/USA/UT-UPHL-201131371/2020\|EPI_ISL_623172\| | ···················· | ···················· | ····················· | ···················· | ···················· | ···················· |
| P | 1976 hCoV-19/USA/UT-UPHL-201130046/2020\|EPI_ISL_623174\| | ···················· | ···················· | ····················· | ···················· | ···················· | ···················· |
| P | 1977 hCoV-19/USA/UT-UPHL-201122069/2020\|EPI_ISL_623175\| | ···················· | ···················· | ····················· | ···················· | ···················· | ···················· |
| P | 1978 hCoV-19/USA/UT-UPHL-201103815/2020\|EPI_ISL_623176\| | ···················· | ···················· | ····················· | ···················· | ···················· | ···················· |
| P | 1979 hCoV-19/USA/UT-UPHL-201132611/2020\|EPI_ISL_623177\| | ···················· | ···················· | ····················· | ···················· | ···················· | ···················· |
| P | 1980 hCoV-19/USA/UT-UPHL-201129311/2020\|EPI_ISL_623178\| | ···················· | ···················· | ····················· | ···················· | ···················· | ···················· |
| P | 1981 hCoV-19/USA/UT-UPHL-201109489/2020\|EPI_ISL_623179\| | ···················· | ···················· | ····················· | ···················· | ···················· | ···················· |
| P | 1982 hCoV-19/England/ALDP-AA45D8/2020\|EPI_ISL_624439\|20 | ···················· | ·A·················· | ····················· | ···················· | ···················· | ···A················ |
| P | 1983 hCoV-19/England/ALDP-AA3C2F/2020\|EPI_ISL_624440\|20 | ···················· | ···················· | ····················· | ···················· | ···················· | ···················· |
| P | 1984 hCoV-19/England/ALDP-AA4453/2020\|EPI_ISL_624441\|20 | ···················· | ···················· | ····················· | ···················· | ···················· | ···················· |
| P | 1985 hCoV-19/England/ALDP-AA3B8C/2020\|EPI_ISL_624442\|20 | ···················· | ···················· | ····················· | ···················· | ···················· | ···················· |
| P | 1986 hCoV-19/England/ALDP-AA43FC/2020\|EPI_ISL_624443\|20 | ···················· | ···················· | ····················· | ···················· | ···················· | ···················· |
| P | 1987 hCoV-19/England/ALDP-AA3B7D/2020\|EPI_ISL_624444\|20 | ···················· | ···················· | ····················· | ···················· | ···················· | ···················· |
| P | 1988 hCoV-19/England/ALDP-AA4356/2020\|EPI_ISL_624446\|20 | ···················· | ···················· | ····················· | ···················· | ···················· | ···················· |
| P | 1989 hCoV-19/England/ALDP-AA2385/2020\|EPI_ISL_624448\|20 | ···················· | ···················· | ····················· | ···················· | ···················· | ···················· |
| P | 1990 hCoV-19/England/ALDP-AA1CD6/2020\|EPI_ISL_624449\|20 | ···················· | ···················· | ····················· | ···················· | ···················· | ···················· |
| P | 1991 hCoV-19/England/ALDP-AA3BC8/2020\|EPI_ISL_624450\|20 | ···················· | ···················· | ····················· | ···················· | ···················· | ···················· |
| P | 1992 hCoV-19/England/ALDP-AA3D4A/2020\|EPI_ISL_624451\|20 | ···················· | ···················· | ····················· | ···················· | ···················· | ···················· |
| P | 1993 hCoV-19/England/ALDP-AA47D2/2020\|EPI_ISL_624453\|20 | ···················· | ···················· | ····················· | ···················· | ···················· | ···················· |
| P | 1994 hCoV-19/England/ALDP-AA240A/2020\|EPI_ISL_624454\|20 | ···················· | ···················· | ····················· | ···················· | ···················· | ···················· |
| P | 1995 hCoV-19/England/ALDP-AA2358/2020\|EPI_ISL_624455\|20 | ···················· | ···················· | ····················· | ···················· | ···················· | ···················· |
| P | 1996 hCoV-19/England/ALDP-AA1562/2020\|EPI_ISL_624456\|20 | ···················· | ···················· | ····················· | ···················· | ···················· | ···················· |
| P | 1997 hCoV-19/England/ALDP-AA21C7/2020\|EPI_ISL_624457\|20 | ···················· | ···················· | ····················· | ···················· | ···················· | ···················· |
| P | 1998 hCoV-19/England/ALDP-AA108F/2020\|EPI_ISL_624458\|20 | ···················· | ···················· | ····················· | ···················· | ···················· | ···················· |
| P | 1999 hCoV-19/England/ALDP-AA27F2/2020\|EPI_ISL_624459\|20 | ···················· | ···················· | ····················· | ···················· | ···················· | ···················· |
| P | 2000 hCoV-19/England/ALDP-AA4857/2020\|EPI_ISL_624460\|20 | ···················· | ···················· | ····················· | ···················· | ···················· | ···················· |
| P | 2001 hCoV-19/England/ALDP-AA1A36/2020\|EPI_ISL_624461\|20 | ···················· | ···················· | ····················· | ···················· | ···················· | ···················· |
| P | 2002 hCoV-19/England/ALDP-AA3E47/2020\|EPI_ISL_624462\|20 | ···················· | ···················· | ····················· | ···················· | ···················· | ···················· |
| P | 2003 hCoV-19/England/ALDP-AA42B3/2020\|EPI_ISL_624463\|20 | ···················· | ···················· | ····················· | ···················· | ···················· | ···················· |
| P | 2004 hCoV-19/England/ALDP-AA2006/2020\|EPI_ISL_624464\|20 | ···················· | ···················· | ····················· | ···················· | ···················· | ···················· |
| P | 2005 hCoV-19/England/ALDP-AA45F6/2020\|EPI_ISL_624465\|20 | ···················· | ···················· | ····················· | ···················· | ···················· | ···················· |
| P | 2006 hCoV-19/England/ALDP-AA1535/2020\|EPI_ISL_624466\|20 | ···················· | ···················· | ····················· | ···················· | ···················· | ···················· |
| P | 2007 hCoV-19/England/ALDP-AA21B8/2020\|EPI_ISL_624467\|20 | ···················· | ···················· | ····················· | ···················· | ···················· | ···················· |
| P | 2008 hCoV-19/England/ALDP-AA1052/2020\|EPI_ISL_624468\|20 | ···················· | ···················· | ····················· | ···················· | ···················· | ···················· |
| P | 2009 hCoV-19/England/ALDP-AA20AC/2020\|EPI_ISL_624469\|20 | ···················· | ···················· | ····················· | ···················· | ···················· | ···················· |
| P | 2010 hCoV-19/England/ALDP-AA1D00/2020\|EPI_ISL_624470\|20 | ···················· | ···················· | ····················· | ···················· | ···················· | ···················· |
| P | 2011 hCoV-19/England/ALDP-AA4BF4/2020\|EPI_ISL_624471\|20 | ···················· | ···················· | ····················· | ···················· | ···················· | ···················· |
| P | 2012 hCoV-19/England/ALDP-AA45AB/2020\|EPI_ISL_624472\|20 | ···················· | ···················· | ····················· | ···················· | ···················· | ···················· |
| P | 2013 hCoV-19/England/ALDP-AA3D2C/2020\|EPI_ISL_624473\|20 | ···················· | ···················· | ····················· | ···················· | ···················· | ···················· |
| P | 2014 hCoV-19/England/ALDP-AA3A07/2020\|EPI_ISL_624474\|20 | ···················· | ·A·······A·········· | ····················· | ···················· | ···················· | ···················· |
| P | 2015 hCoV-19/England/ALDP-AA15BD/2020\|EPI_ISL_624475\|20 | ···················· | ···················· | ····················· | ···················· | ···················· | ···················· |
| P | 2016 hCoV-19/Wales/ALDP-AA47F0/2020\|EPI_ISL_624476\|2020 | ···················· | ···················· | ····················· | ···················· | ···················· | ···················· |
| P | 2017 hCoV-19/England/ALDP-AA1007/2020\|EPI_ISL_624477\|20 | ···················· | ·A·················· | ····················· | ···················· | ···················· | ···················· |
| P | 2018 hCoV-19/England/ALDP-AA3B22/2020\|EPI_ISL_624478\|20 | ···················· | ···················· | ····················· | ···················· | ···················· | ···················· |
| P | 2019 hCoV-19/England/ALDP-AA4C1F/2020\|EPI_ISL_624479\|20 | ···················· | ···················· | ····················· | ···················· | ···················· | ···················· |
| P | 2020 hCoV-19/Denmark/DCGC-8526/2020\|EPI_ISL_625770\|2020 | ···················· | ···················· | ····················· | ···················· | ···················· | ···················· |
| P | 2021 hCoV-19/Denmark/DCGC-8544/2020\|EPI_ISL_625771\|2020 | ···················· | ···················· | ····················· | ···················· | ···················· | ···················· |
| P | 2022 hCoV-19/Denmark/DCGC-8490/2020\|EPI_ISL_625772\|2020 | ···················· | ···················· | ····················· | ···················· | ···C················ | ···················· |
| P | 2023 hCoV-19/Denmark/DCGC-8506/2020\|EPI_ISL_625773\|2020 | ···················· | ···················· | ····················· | ···················· | ···················· | ···················· |
| P | 2024 hCoV-19/Denmark/DCGC-8546/2020\|EPI_ISL_625774\|2020 | ···················· | ···················· | ····················· | ···················· | ···················· | ···················· |
| P | 2025 hCoV-19/Denmark/DCGC-8582/2020\|EPI_ISL_625775\|2020 | ···················· | ···················· | ····················· | ···················· | ···················· | ···················· |
| P | 2026 hCoV-19/Denmark/DCGC-8568/2020\|EPI_ISL_625776\|2020 | ···················· | ···················· | ····················· | ···················· | ···················· | ···················· |
| P | 2027 hCoV-19/Denmark/DCGC-8630/2020\|EPI_ISL_625778\|2020 | ···················· | ···················· | ····················· | ···················· | ···················· | ···················· |
| P | 2028 hCoV-19/Denmark/DCGC-8642/2020\|EPI_ISL_625779\|2020 | ···················· | ···················· | ····················· | ···················· | ···················· | ···················· |
| P | 2029 hCoV-19/Denmark/DCGC-8620/2020\|EPI_ISL_625780\|2020 | ···················· | ···················· | ····················· | ···················· | ···················· | ···················· |
| P | 2030 hCoV-19/Denmark/DCGC-8632/2020\|EPI_ISL_625781\|2020 | ···················· | ···················· | ····················· | ···················· | ···················· | ···················· |
| P | 2031 hCoV-19/Denmark/DCGC-8629/2020\|EPI_ISL_625782\|2020 | ···················· | ···················· | ····················· | ···················· | ···················· | ···················· |
| P | 2032 hCoV-19/Denmark/DCGC-8654/2020\|EPI_ISL_625783\|2020 | ···················· | ···················· | ·········T··········· | ···················· | ···················· | ···················· |
| P | 2033 hCoV-19/Denmark/DCGC-8700/2020\|EPI_ISL_625784\|2020 | ···················· | ···················· | ····················· | ···················· | ···················· | ···················· |
| P | 2034 hCoV-19/Denmark/DCGC-8707/2020\|EPI_ISL_625785\|2020 | ···················· | ···················· | ····················· | ···················· | ···················· | ···················· |
| P | 2035 hCoV-19/Denmark/DCGC-8738/2020\|EPI_ISL_625786\|2020 | ···················· | ···················· | ····················· | ···················· | ···················· | ···················· |

| | | | | | | | | |
|---|---|---|---|---|---|---|---|---|
| P | 2036 | hCoV-19/Denmark/DCGC-8688/2020\|EPI_ISL_625787\|2020 | ···················· | ···················· | ····················· | ···················· | ···················· | ···················· |
| P | 2037 | hCoV-19/Denmark/DCGC-8739/2020\|EPI_ISL_625788\|2020 | ···················· | ···················· | ····················· | ···················· | ···················· | ···················· |
| P | 2038 | hCoV-19/Denmark/DCGC-8689/2020\|EPI_ISL_625789\|2020 | ···················· | ···················· | ····················· | ···················· | ···················· | ···················· |
| P | 2039 | hCoV-19/Denmark/DCGC-8709/2020\|EPI_ISL_625790\|2020 | ···················· | ···················· | ····················· | ···················· | ···················· | ···················· |
| P | 2040 | hCoV-19/Denmark/DCGC-8740/2020\|EPI_ISL_625791\|2020 | ···················· | ···················· | ····················· | ···················· | ···················· | ···················· |
| P | 2041 | hCoV-19/Denmark/DCGC-8736/2020\|EPI_ISL_625792\|2020 | ···················· | ···················· | ····················· | ···················· | ···················· | ···················· |
| P | 2042 | hCoV-19/Denmark/DCGC-8681/2020\|EPI_ISL_625793\|2020 | ···················· | ···················· | ····················· | ···················· | ···················· | ···················· |
| P | 2043 | hCoV-19/Denmark/DCGC-8694/2020\|EPI_ISL_625794\|2020 | ···················· | ···················· | ····················· | ···················· | ···················· | ···················· |
| P | 2044 | hCoV-19/Denmark/DCGC-8699/2020\|EPI_ISL_625795\|2020 | ···················· | ···················· | ····················· | ···················· | ···················· | ···················· |
| P | 2045 | hCoV-19/Denmark/DCGC-8722/2020\|EPI_ISL_625796\|2020 | ···················· | ···················· | ····················· | ···················· | ···················· | ···················· |
| P | 2046 | hCoV-19/Denmark/DCGC-8728/2020\|EPI_ISL_625797\|2020 | ···················· | ···················· | ····················· | ···················· | ···················· | ···················· |
| P | 2047 | hCoV-19/Denmark/DCGC-8737/2020\|EPI_ISL_625798\|2020 | ···················· | ···················· | ····················· | ···················· | ···················· | ···················· |
| P | 2048 | hCoV-19/Denmark/DCGC-8763/2020\|EPI_ISL_625799\|2020 | ···················· | ···················· | ····················· | ···················· | ···················· | ···················· |
| P | 2049 | hCoV-19/Denmark/DCGC-3189/2020\|EPI_ISL_616012\|2020 | ···················· | ···················· | ····················· | ···················· | ···················· | ···················· |
| P | 2050 | hCoV-19/Denmark/DCGC-8163/2020\|EPI_ISL_617230\|2020 | ···················· | ···················· | ····················· | ···················· | ···················· | ···················· |
| P | 2051 | hCoV-19/Denmark/DCGC-8114/2020\|EPI_ISL_617231\|2020 | ···················· | ·A·················· | ····················· | ···················· | ···················· | ···················· |
| P | 2052 | hCoV-19/Denmark/DCGC-8142/2020\|EPI_ISL_617232\|2020 | ···················· | ···················· | ····················· | ···················· | ···················· | ···················· |
| P | 2053 | hCoV-19/Denmark/DCGC-8150/2020\|EPI_ISL_617233\|2020 | ···················· | ···················· | ····················· | ···················· | ···················· | ···················· |
| P | 2054 | hCoV-19/Denmark/DCGC-8158/2020\|EPI_ISL_617234\|2020 | ···················· | ···················· | ····················· | ···················· | ···················· | ···················· |
| P | 2055 | hCoV-19/Denmark/DCGC-8176/2020\|EPI_ISL_617235\|2020 | ···················· | ···················· | ····················· | ···················· | ···················· | ···················· |
| P | 2056 | hCoV-19/Denmark/DCGC-8115/2020\|EPI_ISL_617236\|2020 | ···················· | ···················· | ····················· | ···················· | ···················· | ···················· |
| P | 2057 | hCoV-19/Denmark/DCGC-8135/2020\|EPI_ISL_617237\|2020 | ···················· | ···················· | ····················· | ···················· | ···················· | ···················· |
| P | 2058 | hCoV-19/Denmark/DCGC-8143/2020\|EPI_ISL_617238\|2020 | ···················· | ···················· | ····················· | ···················· | ···················· | ···················· |
| P | 2059 | hCoV-19/Denmark/DCGC-8159/2020\|EPI_ISL_617239\|2020 | ···················· | ···················· | ····················· | ···················· | ···················· | ···················· |
| P | 2060 | hCoV-19/Denmark/DCGC-8165/2020\|EPI_ISL_617240\|2020 | ···················· | ···················· | ····················· | ···················· | ···················· | ···················· |
| P | 2061 | hCoV-19/Denmark/DCGC-8177/2020\|EPI_ISL_617241\|2020 | ···················· | ···················· | ····················· | ···················· | ···················· | ···················· |
| P | 2062 | hCoV-19/Denmark/DCGC-8116/2020\|EPI_ISL_617242\|2020 | ···················· | A··················· | ····················· | ···················· | ···················· | ···················· |
| P | 2063 | hCoV-19/Denmark/DCGC-8124/2020\|EPI_ISL_617243\|2020 | ···················· | ····A··············· | ····················· | ···················· | ···················· | ···················· |
| P | 2064 | hCoV-19/Denmark/DCGC-8130/2020\|EPI_ISL_617244\|2020 | ···················· | ···················· | ····················· | ···················· | ···················· | ···················· |
| P | 2065 | hCoV-19/Denmark/DCGC-8144/2020\|EPI_ISL_617245\|2020 | ···················· | ···················· | ····················· | ···················· | ···················· | ···················· |
| P | 2066 | hCoV-19/Denmark/DCGC-8160/2020\|EPI_ISL_617246\|2020 | ···················· | ···················· | ····················· | ···················· | ···················· | ···················· |
| P | 2067 | hCoV-19/Denmark/DCGC-8166/2020\|EPI_ISL_617247\|2020 | ···················· | ···················· | ····················· | ···················· | ···················· | ···················· |
| P | 2068 | hCoV-19/Denmark/DCGC-8171/2020\|EPI_ISL_617248\|2020 | ···················· | ···················· | ····················· | ···················· | ···················· | ···················· |
| P | 2069 | hCoV-19/Denmark/DCGC-8185/2020\|EPI_ISL_617249\|2020 | ···················· | ···················· | ····················· | ···················· | ···················· | ···················· |
| P | 2070 | hCoV-19/Denmark/DCGC-8198/2020\|EPI_ISL_617250\|2020 | ···················· | ···················· | ····················· | ···················· | ···················· | ···················· |
| P | 2071 | hCoV-19/Denmark/DCGC-8209/2020\|EPI_ISL_617251\|2020 | ···················· | ···················· | ····················· | ···················· | ···················· | ···················· |
| P | 2072 | hCoV-19/Denmark/DCGC-8213/2020\|EPI_ISL_617252\|2020 | ···················· | ···················· | ····················· | ···················· | ···················· | ···················· |
| P | 2073 | hCoV-19/Denmark/DCGC-8223/2020\|EPI_ISL_617253\|2020 | ···················· | ···················· | ····················· | ···················· | ···················· | ···················· |
| P | 2074 | hCoV-19/Denmark/DCGC-8231/2020\|EPI_ISL_617254\|2020 | ···················· | ···················· | ····················· | ···················· | ···················· | ···················· |
| P | 2075 | hCoV-19/Denmark/DCGC-8249/2020\|EPI_ISL_617255\|2020 | ···················· | ···················· | ····················· | ···················· | ···················· | ···················· |
| P | 2076 | hCoV-19/Denmark/DCGC-8199/2020\|EPI_ISL_617256\|2020 | ···················· | ···················· | ····················· | ···················· | ···················· | ···················· |
| P | 2077 | hCoV-19/Denmark/DCGC-8210/2020\|EPI_ISL_617257\|2020 | ···················· | ···················· | ····················· | ···················· | ···················· | ···················· |
| P | 2078 | hCoV-19/Denmark/DCGC-8214/2020\|EPI_ISL_617258\|2020 | ···················· | ···················· | ····················· | ···················· | ···················· | ···················· |
| P | 2079 | hCoV-19/Denmark/DCGC-8224/2020\|EPI_ISL_617259\|2020 | ···················· | ···················· | ····················· | ···················· | ···················· | ···················· |
| P | 2080 | hCoV-19/Denmark/DCGC-8225/2020\|EPI_ISL_617262\|2020 | ···················· | ···················· | ····················· | ···················· | ···················· | ···················· |
| P | 2081 | hCoV-19/Denmark/DCGC-8257/2020\|EPI_ISL_617263\|2020 | ···················· | ···················· | ····················· | ···················· | ···················· | ···················· |
| P | 2082 | hCoV-19/Denmark/DCGC-8204/2020\|EPI_ISL_617266\|2020 | ···················· | ···················· | ····················· | ···················· | ···················· | ···················· |
| P | 2083 | hCoV-19/Denmark/DCGC-8226/2020\|EPI_ISL_617267\|2020 | ···················· | ···················· | ····················· | ···················· | ···················· | ···················· |
| P | 2084 | hCoV-19/Denmark/DCGC-8234/2020\|EPI_ISL_617268\|2020 | ···················· | ···················· | ····················· | ···················· | ···················· | ···················· |
| P | 2085 | hCoV-19/Denmark/DCGC-8241/2020\|EPI_ISL_617269\|2020 | ···················· | ···················· | ····················· | ···················· | ···················· | ···················· |
| P | 2086 | hCoV-19/Denmark/DCGC-8194/2020\|EPI_ISL_617270\|2020 | ···················· | ···················· | ····················· | ···················· | ···················· | ···················· |
| P | 2087 | hCoV-19/Denmark/DCGC-8246/2020\|EPI_ISL_617271\|2020 | ···················· | ···················· | ····················· | ···················· | ···················· | ···················· |
| P | 2088 | hCoV-19/Denmark/DCGC-8258/2020\|EPI_ISL_617272\|2020 | ···················· | ·A·················· | ····················· | ···················· | ···················· | ···················· |
| P | 2089 | hCoV-19/Denmark/DCGC-8228/2020\|EPI_ISL_617273\|2020 | ···················· | ···················· | ····················· | ···················· | ···················· | ···················· |
| P | 2090 | hCoV-19/Denmark/DCGC-8236/2020\|EPI_ISL_617274\|2020 | ···················· | ···················· | ····················· | ···················· | ···················· | ···················· |
| P | 2091 | hCoV-19/Denmark/DCGC-8196/2020\|EPI_ISL_617275\|2020 | ···················· | ···················· | ····················· | ···················· | ···················· | ···················· |
| P | 2092 | hCoV-19/Denmark/DCGC-8207/2020\|EPI_ISL_617276\|2020 | ···················· | ···················· | ····················· | ···················· | ···················· | ···················· |
| P | 2093 | hCoV-19/Denmark/DCGC-8217/2020\|EPI_ISL_617277\|2020 | ···················· | ···················· | ····················· | ···················· | ···················· | ···················· |
| P | 2094 | hCoV-19/Denmark/DCGC-8229/2020\|EPI_ISL_617278\|2020 | ···················· | ···················· | ····················· | ···················· | ···················· | ···················· |
| P | 2095 | hCoV-19/Denmark/DCGC-8197/2020\|EPI_ISL_617279\|2020 | ···················· | ···················· | ····················· | ···················· | ···················· | ···················· |
| P | 2096 | hCoV-19/Denmark/DCGC-8212/2020\|EPI_ISL_617280\|2020 | ···················· | ···················· | ····················· | ···················· | ···················· | ···················· |
| P | 2097 | hCoV-19/Denmark/DCGC-8230/2020\|EPI_ISL_617281\|2020 | ···················· | ···················· | ····················· | ···················· | ···················· | ···················· |
| P | 2098 | hCoV-19/Denmark/DCGC-8238/2020\|EPI_ISL_617282\|2020 | ···················· | ···················· | ····················· | ···················· | ···················· | ···················· |
| P | 2099 | hCoV-19/Denmark/DCGC-8254/2020\|EPI_ISL_617283\|2020 | ···················· | ···················· | ····················· | ···················· | ···················· | ···················· |
| P | 2100 | hCoV-19/Denmark/DCGC-1674/2020\|EPI_ISL_617284\|2020 | ···················· | ···················· | ····················· | ···················· | ···················· | ···················· |
| P | 2101 | hCoV-19/Denmark/DCGC-8259/2020\|EPI_ISL_617285\|2020 | ···················· | ·A····A············· | ····················· | ···················· | ···················· | ···················· |
| P | 2102 | hCoV-19/Denmark/DCGC-8266/2020\|EPI_ISL_617286\|2020 | ···················· | ·A····A············· | ····················· | ···················· | ···················· | ···················· |
| P | 2103 | hCoV-19/Denmark/DCGC-8274/2020\|EPI_ISL_617287\|2020 | ···················· | ···················· | ····················· | ···················· | ···················· | ···················· |
| P | 2104 | hCoV-19/Denmark/DCGC-8285/2020\|EPI_ISL_617288\|2020 | ···················· | ·A·················· | ····················· | ···················· | ···················· | ···················· |
| P | 2105 | hCoV-19/Denmark/DCGC-8290/2020\|EPI_ISL_617289\|2020 | ···················· | ···················· | ····················· | ···················· | ···················· | ···················· |
| P | 2106 | hCoV-19/Denmark/DCGC-8298/2020\|EPI_ISL_617290\|2020 | ···················· | ···················· | ····················· | ···················· | ···················· | ···················· |
| P | 2107 | hCoV-19/Denmark/DCGC-8323/2020\|EPI_ISL_617291\|2020 | ···················· | ···················· | ····················· | ···················· | ···················· | ···················· |
| P | 2108 | hCoV-19/Denmark/DCGC-8260/2020\|EPI_ISL_617292\|2020 | ···················· | ····A··············· | ····················· | ···················· | ···················· | ···················· |
| P | 2109 | hCoV-19/Denmark/DCGC-8267/2020\|EPI_ISL_617293\|2020 | ···················· | ···················· | ····················· | ···················· | ···················· | ···················· |
| P | 2110 | hCoV-19/Denmark/DCGC-8312/2020\|EPI_ISL_617294\|2020 | ···················· | ···················· | ····················· | ···················· | ···················· | ···················· |
| P | 2111 | hCoV-19/Denmark/DCGC-8316/2020\|EPI_ISL_617295\|2020 | ···················· | ···················· | ····················· | ···················· | ···················· | ···················· |
| P | 2112 | hCoV-19/Denmark/DCGC-8268/2020\|EPI_ISL_617296\|2020 | ···················· | ···················· | ····················· | ···················· | ···················· | ···················· |
| P | 2113 | hCoV-19/Denmark/DCGC-8292/2020\|EPI_ISL_617297\|2020 | ···················· | ···················· | ····················· | ···················· | ···················· | ···················· |
| P | 2114 | hCoV-19/Denmark/DCGC-8300/2020\|EPI_ISL_617298\|2020 | ···················· | ···················· | ····················· | ···················· | ···················· | ···················· |
| P | 2115 | hCoV-19/Denmark/DCGC-8317/2020\|EPI_ISL_617299\|2020 | ···················· | ···················· | ····················· | ···················· | ···················· | ···················· |

| | | | | | | | | |
|---|---|---|---|---|---|---|---|---|
| P | 2116 | hCoV-19/Denmark/DCGC-8324/2020\|EPI_ISL_617300\|2020 | ···················· | ···················· | ····················· | ···················· | ···················· | ···················· |
| P | 2117 | hCoV-19/Denmark/DCGC-8331/2020\|EPI_ISL_617301\|2020 | ···················· | ···················· | ····················· | ···················· | ···················· | ···················· |
| P | 2118 | hCoV-19/Denmark/DCGC-8293/2020\|EPI_ISL_617302\|2020 | ···················· | ···················· | ····················· | ···················· | ···················· | ···················· |
| P | 2119 | hCoV-19/Denmark/DCGC-8301/2020\|EPI_ISL_617303\|2020 | ···················· | ···················· | ····················· | ···················· | ···················· | ···················· |
| P | 2120 | hCoV-19/Denmark/DCGC-8307/2020\|EPI_ISL_617304\|2020 | ···················· | ···················· | ····················· | ···················· | ···················· | ···················· |
| P | 2121 | hCoV-19/Denmark/DCGC-8313/2020\|EPI_ISL_617305\|2020 | ···················· | ···················· | ····················· | ···················· | ···················· | ···················· |
| P | 2122 | hCoV-19/Denmark/DCGC-8332/2020\|EPI_ISL_617306\|2020 | ···················· | ···················· | ····················· | ···················· | ···················· | ···················· |
| P | 2123 | hCoV-19/Denmark/DCGC-8262/2020\|EPI_ISL_617307\|2020 | ···················· | ···················· | ····················· | ···················· | ···················· | ···················· |
| P | 2124 | hCoV-19/Denmark/DCGC-8276/2020\|EPI_ISL_617308\|2020 | ···················· | ···················· | ····················· | ···················· | ···················· | ···················· |
| P | 2125 | hCoV-19/Denmark/DCGC-8282/2020\|EPI_ISL_617309\|2020 | ···················· | ···················· | ····················· | ···················· | ···················· | ···················· |
| P | 2126 | hCoV-19/Denmark/DCGC-8401/2020\|EPI_ISL_617342\|2020 | ···················· | ···················· | ····················· | ···················· | ···················· | ···················· |
| P | 2127 | hCoV-19/Denmark/DCGC-7671/2020\|EPI_ISL_618560\|2020 | ···················· | ···················· | ····················· | ···················· | ···················· | ···················· |
| P | 2128 | hCoV-19/Denmark/DCGC-7675/2020\|EPI_ISL_618561\|2020 | ···················· | ···················· | ····················· | ···················· | ···················· | ···················· |
| P | 2129 | hCoV-19/Denmark/DCGC-7681/2020\|EPI_ISL_618562\|2020 | ···················· | ·········N·········· | ····················· | ···················· | ···················· | ···················· |
| P | 2130 | hCoV-19/Denmark/DCGC-7699/2020\|EPI_ISL_618563\|2020 | ···················· | ···················· | ····················· | ···················· | ···················· | ···················· |
| P | 2131 | hCoV-19/Denmark/DCGC-7709/2020\|EPI_ISL_618564\|2020 | ···················· | ···················· | ····················· | ···················· | ···················· | ···················· |
| P | 2132 | hCoV-19/Denmark/DCGC-7716/2020\|EPI_ISL_618565\|2020 | ···················· | ···················· | ····················· | ···················· | ···················· | ···················· |
| P | 2133 | hCoV-19/Denmark/DCGC-7656/2020\|EPI_ISL_618566\|2020 | ···················· | ·A·················· | ····················· | ···················· | ···················· | ··A················· |
| P | 2134 | hCoV-19/Denmark/DCGC-7664/2020\|EPI_ISL_618567\|2020 | ···················· | ···················· | ····················· | ···················· | ···················· | ···················· |
| P | 2135 | hCoV-19/Denmark/DCGC-7672/2020\|EPI_ISL_618568\|2020 | ···················· | ···················· | ····················· | ···················· | ···················· | ···················· |
| P | 2136 | hCoV-19/Denmark/DCGC-7676/2020\|EPI_ISL_618569\|2020 | ···················· | ···················· | ····················· | ···················· | ···················· | ···················· |
| P | 2137 | hCoV-19/Denmark/DCGC-7682/2020\|EPI_ISL_618570\|2020 | ···················· | ·A·················· | ····················· | ···················· | ···················· | ··A················· |
| P | 2138 | hCoV-19/Denmark/DCGC-7705/2020\|EPI_ISL_618573\|2020 | ···················· | ···················· | ····················· | ···················· | ···················· | ···················· |
| P | 2139 | hCoV-19/Denmark/DCGC-7710/2020\|EPI_ISL_618574\|2020 | ···················· | ···················· | ····················· | ···················· | ···················· | ···················· |
| P | 2140 | hCoV-19/Denmark/DCGC-7717/2020\|EPI_ISL_618575\|2020 | ···················· | ···················· | ····················· | ···················· | ···················· | ···················· |
| P | 2141 | hCoV-19/Denmark/DCGC-7657/2020\|EPI_ISL_618576\|2020 | ···················· | ···················· | ····················· | ···················· | ···················· | ···················· |
| P | 2142 | hCoV-19/Denmark/DCGC-7665/2020\|EPI_ISL_618577\|2020 | ···················· | ···················· | ····················· | ···················· | ···················· | ···················· |
| P | 2143 | hCoV-19/Denmark/DCGC-7673/2020\|EPI_ISL_618578\|2020 | ···················· | ·A·················· | ····················· | ···················· | ···················· | ···················· |
| P | 2144 | hCoV-19/Denmark/DCGC-7686/2020\|EPI_ISL_618579\|2020 | ···················· | ···················· | ····················· | ···················· | ···················· | ···················· |
| P | 2145 | hCoV-19/Denmark/DCGC-7706/2020\|EPI_ISL_618580\|2020 | ···················· | ···················· | ····················· | ···················· | ···················· | ···················· |
| P | 2146 | hCoV-19/Denmark/DCGC-7711/2020\|EPI_ISL_618581\|2020 | ···················· | ···················· | ····················· | ···················· | ······T············· | ···················· |
| P | 2147 | hCoV-19/Denmark/DCGC-7718/2020\|EPI_ISL_618582\|2020 | ···················· | ·A·················· | ····················· | ···················· | ···················· | ··A················· |
| P | 2148 | hCoV-19/Denmark/DCGC-7658/2020\|EPI_ISL_618583\|2020 | ···················· | ···················· | ····················· | ···················· | ···················· | ···················· |
| P | 2149 | hCoV-19/Denmark/DCGC-7666/2020\|EPI_ISL_618584\|2020 | ···················· | ···················· | ····················· | ···················· | ···················· | ···················· |
| P | 2150 | hCoV-19/Denmark/DCGC-7677/2020\|EPI_ISL_618585\|2020 | ···················· | ···················· | ····················· | ···················· | ···················· | ···················· |
| P | 2151 | hCoV-19/Denmark/DCGC-7687/2020\|EPI_ISL_618586\|2020 | ···················· | ···················· | ····················· | ···················· | ···················· | ···················· |
| P | 2152 | hCoV-19/Denmark/DCGC-7690/2020\|EPI_ISL_618587\|2020 | ···················· | ···················· | ····················· | ···················· | ···················· | ···················· |
| P | 2153 | hCoV-19/Denmark/DCGC-7659/2020\|EPI_ISL_618593\|2020 | ···················· | ···················· | ····················· | ···················· | ···················· | ···················· |
| P | 2154 | hCoV-19/Denmark/DCGC-7667/2020\|EPI_ISL_618594\|2020 | ···················· | ···················· | ····················· | ···················· | ···················· | ···················· |
| P | 2155 | hCoV-19/Denmark/DCGC-7691/2020\|EPI_ISL_618597\|2020 | ···················· | ···················· | ····················· | ···················· | ···················· | ···················· |
| P | 2156 | hCoV-19/Denmark/DCGC-7695/2020\|EPI_ISL_618598\|2020 | ···················· | ···················· | ····················· | ···················· | ···················· | ···················· |
| P | 2157 | hCoV-19/Denmark/DCGC-7713/2020\|EPI_ISL_618600\|2020 | ···················· | ···················· | ····················· | ···················· | ···················· | ···················· |
| P | 2158 | hCoV-19/Denmark/DCGC-7720/2020\|EPI_ISL_618601\|2020 | ···················· | ···················· | ····················· | ···················· | ···················· | ···················· |
| P | 2159 | hCoV-19/Denmark/DCGC-7660/2020\|EPI_ISL_618602\|2020 | ···················· | ···················· | ····················· | ···················· | ······T············· | ···················· |
| P | 2160 | hCoV-19/Denmark/DCGC-7668/2020\|EPI_ISL_618603\|2020 | ···················· | ·A·················· | ····················· | ···················· | ···················· | ··A················· |
| P | 2161 | hCoV-19/Denmark/DCGC-7688/2020\|EPI_ISL_618604\|2020 | ···················· | ···················· | ····················· | ···················· | ···················· | ···················· |
| P | 2162 | hCoV-19/Denmark/DCGC-7692/2020\|EPI_ISL_618605\|2020 | ···················· | ···················· | ····················· | ···················· | ···················· | ···················· |
| P | 2163 | hCoV-19/Denmark/DCGC-7696/2020\|EPI_ISL_618606\|2020 | ···················· | ···················· | ····················· | ···················· | ···················· | ···················· |
| P | 2164 | hCoV-19/Denmark/DCGC-7661/2020\|EPI_ISL_618607\|2020 | ···················· | ···················· | ····················· | ···················· | ······T············· | ···················· |
| P | 2165 | hCoV-19/Denmark/DCGC-7669/2020\|EPI_ISL_618608\|2020 | ···················· | ···················· | ····················· | ···················· | ···················· | ···················· |
| P | 2166 | hCoV-19/Denmark/DCGC-1581/2020\|EPI_ISL_618609\|2020 | ···················· | ···················· | ····················· | ···················· | ···················· | ···················· |
| P | 2167 | hCoV-19/Denmark/DCGC-7679/2020\|EPI_ISL_618610\|2020 | ···················· | ···················· | ····················· | ···················· | ···················· | ···················· |
| P | 2168 | hCoV-19/Denmark/DCGC-7683/2020\|EPI_ISL_618611\|2020 | ···················· | ···················· | ····················· | ···················· | ···················· | ···················· |
| P | 2169 | hCoV-19/Denmark/DCGC-7689/2020\|EPI_ISL_618612\|2020 | ···················· | ···················· | ····················· | ···················· | ···················· | ···················· |
| P | 2170 | hCoV-19/Denmark/DCGC-7697/2020\|EPI_ISL_618613\|2020 | ···················· | ···················· | ····················· | ···················· | ···················· | ···················· |
| P | 2171 | hCoV-19/Denmark/DCGC-7703/2020\|EPI_ISL_618614\|2020 | ···················· | ···················· | ····················· | ···················· | ···················· | ···················· |
| P | 2172 | hCoV-19/Denmark/DCGC-7714/2020\|EPI_ISL_618615\|2020 | ···················· | ···················· | ····················· | ···················· | ···················· | ···················· |
| P | 2173 | hCoV-19/Denmark/DCGC-7662/2020\|EPI_ISL_618616\|2020 | ···················· | ···················· | ····················· | ···················· | ···················· | ···················· |
| P | 2174 | hCoV-19/Denmark/DCGC-7670/2020\|EPI_ISL_618617\|2020 | ···················· | ···················· | ····················· | ···················· | ···················· | ···················· |
| P | 2175 | hCoV-19/Denmark/DCGC-7680/2020\|EPI_ISL_618619\|2020 | ···················· | ···················· | ····················· | ···················· | ···················· | ···················· |
| P | 2176 | hCoV-19/Denmark/DCGC-7684/2020\|EPI_ISL_618620\|2020 | ···················· | ···················· | ····················· | ···················· | ···················· | ···················· |
| P | 2177 | hCoV-19/Denmark/DCGC-7698/2020\|EPI_ISL_618622\|2020 | ···················· | ···················· | ····················· | ···················· | ···················· | ···················· |
| P | 2178 | hCoV-19/Denmark/DCGC-7715/2020\|EPI_ISL_618625\|2020 | ···················· | ···················· | ····················· | ···················· | ···················· | ···················· |
| P | 2179 | hCoV-19/Denmark/DCGC-1582/2020\|EPI_ISL_618626\|2020 | ···················· | ···················· | ····················· | ···················· | ···················· | ···················· |
| P | 2180 | hCoV-19/Denmark/DCGC-1583/2020\|EPI_ISL_618627\|2020 | ···················· | ···················· | ····················· | ···················· | ···················· | ···················· |
| P | 2181 | hCoV-19/Denmark/DCGC-1584/2020\|EPI_ISL_618628\|2020 | ···················· | ···················· | ····················· | ···················· | ···················· | ···················· |
| P | 2182 | hCoV-19/Denmark/DCGC-1585/2020\|EPI_ISL_618629\|2020 | ···················· | ···················· | ····················· | ···················· | ···················· | ···················· |
| P | 2183 | hCoV-19/Denmark/DCGC-1586/2020\|EPI_ISL_618630\|2020 | ···················· | ···················· | ····················· | ···················· | ···················· | ···················· |
| P | 2184 | hCoV-19/Denmark/DCGC-1587/2020\|EPI_ISL_618631\|2020 | ···················· | ···················· | ····················· | ···················· | ···················· | ···················· |
| P | 2185 | hCoV-19/Denmark/DCGC-1589/2020\|EPI_ISL_618633\|2020 | ···················· | ···················· | ····················· | ···················· | ···················· | ···················· |
| P | 2186 | hCoV-19/Denmark/DCGC-1590/2020\|EPI_ISL_618634\|2020 | ···················· | ···················· | ····················· | ···················· | ···················· | ···················· |
| P | 2187 | hCoV-19/Denmark/DCGC-1591/2020\|EPI_ISL_618635\|2020 | ···················· | ···················· | ····················· | ···················· | ···················· | ···················· |
| P | 2188 | hCoV-19/Denmark/DCGC-1593/2020\|EPI_ISL_618637\|2020 | ···················· | ···················· | ····················· | ···················· | ···················· | ···················· |
| P | 2189 | hCoV-19/Denmark/DCGC-1594/2020\|EPI_ISL_618638\|2020 | ···················· | ···················· | ····················· | ···················· | ···················· | ···················· |
| P | 2190 | hCoV-19/Denmark/DCGC-1595/2020\|EPI_ISL_618639\|2020 | ···················· | ···················· | ····················· | ···················· | ···················· | ···················· |
| P | 2191 | hCoV-19/Denmark/DCGC-7788/2020\|EPI_ISL_618673\|2020 | ···················· | ·A·················· | ····················· | ···················· | ···················· | ···················· |
| P | 2192 | hCoV-19/Denmark/DCGC-7043/2020\|EPI_ISL_619891\|2020 | ···················· | ···················· | ····················· | ···················· | ···················· | ···················· |
| P | 2193 | hCoV-19/Denmark/DCGC-7045/2020\|EPI_ISL_619893\|2020 | ···················· | ···················· | ····················· | ···················· | ···················· | ···················· |
| P | 2194 | hCoV-19/Denmark/DCGC-7046/2020\|EPI_ISL_619894\|2020 | ···················· | ···················· | ····················· | ···················· | ···················· | ···················· |
| P | 2195 | hCoV-19/Denmark/DCGC-1644/2020\|EPI_ISL_619895\|2020 | ···················· | ···················· | ····················· | ···················· | ···················· | ···················· |

```
P 2196 hCoV-19/Denmark/DCGC-7048/2020|EPI_ISL_619897|2020 .................... .................... .................... .................... .................... ....................
P 2197 hCoV-19/Denmark/DCGC-7049/2020|EPI_ISL_619898|2020 .................... .................... .................... .................... .................... ....................
P 2198 hCoV-19/Denmark/DCGC-7050/2020|EPI_ISL_619899|2020 .................... .................... .................... .................... .................... ....................
P 2199 hCoV-19/Denmark/DCGC-5642/2020|EPI_ISL_619929|2020 .................... .................... .................... .................... .................... ....................
P 2200 hCoV-19/Denmark/DCGC-5643/2020|EPI_ISL_619930|2020 .................... .................... .................... .................... .................... ....................
P 2201 hCoV-19/Denmark/DCGC-5644/2020|EPI_ISL_619931|2020 .................... .................... .................... .................... .................... ....................
P 2202 hCoV-19/Denmark/DCGC-5645/2020|EPI_ISL_619932|2020 .................... .................... .................... .................... .................... ....................
P 2203 hCoV-19/Denmark/DCGC-5646/2020|EPI_ISL_619933|2020 .................... .................... .................... .................... .................... ....................
P 2204 hCoV-19/Denmark/DCGC-5647/2020|EPI_ISL_619934|2020 .................... .................... .................... .................... .................... ....................
P 2205 hCoV-19/Denmark/DCGC-5648/2020|EPI_ISL_619935|2020 .................... .................... .................... .................... .................... ....................
P 2206 hCoV-19/Denmark/DCGC-5649/2020|EPI_ISL_619936|2020 .................... .................... .................... .................... .................... ....................
P 2207 hCoV-19/Denmark/DCGC-5650/2020|EPI_ISL_619937|2020 .................... .................... .................... .................... .................... ....................
P 2208 hCoV-19/Denmark/DCGC-5651/2020|EPI_ISL_619938|2020 .................... .................... .................... .................... .................... ....................
P 2209 hCoV-19/Denmark/DCGC-5652/2020|EPI_ISL_619939|2020 .................... .................... .................... .................... .................... ....................
P 2210 hCoV-19/Denmark/DCGC-5653/2020|EPI_ISL_619940|2020 .................... .................... .................... .................... .................... ....................
P 2211 hCoV-19/Denmark/DCGC-5654/2020|EPI_ISL_619941|2020 .................... .................... .................... .................... .................... ....................
P 2212 hCoV-19/Denmark/DCGC-5656/2020|EPI_ISL_619943|2020 .................... .................... .................... .................... .................... ....................
P 2213 hCoV-19/Denmark/DCGC-5657/2020|EPI_ISL_619944|2020 .................... .................... .................... .................... .................... ....................
P 2214 hCoV-19/Denmark/DCGC-5658/2020|EPI_ISL_619945|2020 .................... .................... .................... .................... .................... ....................
P 2215 hCoV-19/Denmark/DCGC-5659/2020|EPI_ISL_619946|2020 .................... .................... .................... .................... .................... ....................
P 2216 hCoV-19/Denmark/DCGC-5662/2020|EPI_ISL_619949|2020 .................... .................... .................... .................... .................... ....................
P 2217 hCoV-19/Denmark/DCGC-5663/2020|EPI_ISL_619950|2020 .................... .................... .................... .................... .................... ....................
P 2218 hCoV-19/Denmark/DCGC-5664/2020|EPI_ISL_619951|2020 .................... .................... .................... .................... .................... ....................
P 2219 hCoV-19/Denmark/DCGC-5665/2020|EPI_ISL_619952|2020 .................... .................... .................... .................... .................... ....................
P 2220 hCoV-19/Denmark/DCGC-5667/2020|EPI_ISL_619954|2020 .................... .................... .................... .................... .................... ....................
P 2221 hCoV-19/Denmark/DCGC-5668/2020|EPI_ISL_619955|2020 .................... .................... .................... .................... .................... ....................
P 2222 hCoV-19/Denmark/DCGC-5669/2020|EPI_ISL_619956|2020 .................... .................... .................... .................... .................... ....................
P 2223 hCoV-19/Denmark/DCGC-5670/2020|EPI_ISL_619957|2020 .................... .................... .................... .................... .................... ....................
P 2224 hCoV-19/Denmark/DCGC-5671/2020|EPI_ISL_619958|2020 .................... .................... .................... .................... .................... ....................
P 2225 hCoV-19/Denmark/DCGC-5672/2020|EPI_ISL_619959|2020 .................... .................... .................... .................... .................... ....................
P 2226 hCoV-19/Denmark/DCGC-5673/2020|EPI_ISL_619960|2020 .................... .................... .................... .................... .................... ....................
P 2227 hCoV-19/Denmark/DCGC-5674/2020|EPI_ISL_619961|2020 .................... .................... .................... .................... .................... ....................
P 2228 hCoV-19/Denmark/DCGC-5675/2020|EPI_ISL_619962|2020 .................... .................... .................... .................... .................... ....................
P 2229 hCoV-19/Denmark/DCGC-5677/2020|EPI_ISL_619964|2020 .................... .................... .................... .................... .................... ....................
P 2230 hCoV-19/Denmark/DCGC-5678/2020|EPI_ISL_619965|2020 .................... .................... .................... .................... .................... ....................
P 2231 hCoV-19/Denmark/DCGC-5680/2020|EPI_ISL_619967|2020 .................... .A.................. .................... .................... .................... ....................
P 2232 hCoV-19/Denmark/DCGC-5681/2020|EPI_ISL_619968|2020 .................... .................... .................... .................... .................... ....................
P 2233 hCoV-19/Denmark/DCGC-5682/2020|EPI_ISL_619969|2020 .................... .................... .................... .................... .................... ....................
P 2234 hCoV-19/Denmark/DCGC-5552/2020|EPI_ISL_620877|2020 .................... .................... .................... .................... .................... ....................
P 2235 hCoV-19/Denmark/DCGC-5554/2020|EPI_ISL_620879|2020 .................... .................... .................... .................... .................... ....................
P 2236 hCoV-19/Denmark/DCGC-5555/2020|EPI_ISL_620880|2020 .................... .................... .................... .................... .................... ....................
P 2237 hCoV-19/Denmark/DCGC-5556/2020|EPI_ISL_620881|2020 .................... .................... .................... .................... .................... ....................
P 2238 hCoV-19/Denmark/DCGC-5557/2020|EPI_ISL_620882|2020 .................... .................... .................... .................... .................... ....................
P 2239 hCoV-19/Denmark/DCGC-5558/2020|EPI_ISL_620883|2020 .................... .................... .................... .................... .................... ....................
P 2240 hCoV-19/Denmark/DCGC-5559/2020|EPI_ISL_620884|2020 .................... .................... .................... .................... .................... ....................
P 2241 hCoV-19/Denmark/DCGC-5560/2020|EPI_ISL_620885|2020 .................... .................... .................... .................... .................... ....................
P 2242 hCoV-19/Denmark/DCGC-5561/2020|EPI_ISL_620886|2020 .................... .A.................. .................... .................... .................... ....................
P 2243 hCoV-19/Denmark/DCGC-5562/2020|EPI_ISL_620887|2020 .................... .................... .................... .................... .................... ....................
P 2244 hCoV-19/Denmark/DCGC-5563/2020|EPI_ISL_620888|2020 .................... .................... .................... .................... .................... ....................
P 2245 hCoV-19/Denmark/DCGC-5564/2020|EPI_ISL_620889|2020 .................... .................... .................... .................... .................... ....................
P 2246 hCoV-19/Denmark/DCGC-5565/2020|EPI_ISL_620890|2020 .................... .................... .................... .................... .................... ....................
P 2247 hCoV-19/Denmark/DCGC-5567/2020|EPI_ISL_620892|2020 .................... .................... .................... .................... .................... ....................
P 2248 hCoV-19/Denmark/DCGC-5568/2020|EPI_ISL_620893|2020 .................... .................... .................... .................... .................... ....................
P 2249 hCoV-19/Denmark/DCGC-5569/2020|EPI_ISL_620894|2020 .................... .................... .................... .................... .................... ....................
P 2250 hCoV-19/Denmark/DCGC-5570/2020|EPI_ISL_620895|2020 .................... .................... .................... .................... .................... ....................
P 2251 hCoV-19/Denmark/DCGC-5571/2020|EPI_ISL_620896|2020 .................... .................... .................... .................... ......T............. ....................
P 2252 hCoV-19/Denmark/DCGC-5572/2020|EPI_ISL_620897|2020 .................... .................... .................... .................... .................... ....................
P 2253 hCoV-19/Denmark/DCGC-5573/2020|EPI_ISL_620898|2020 .................... .................... .................... .................... .................... ....................
P 2254 hCoV-19/Denmark/DCGC-5574/2020|EPI_ISL_620899|2020 .................... .................... .................... .................... .................... ....................
P 2255 hCoV-19/Denmark/DCGC-5585/2020|EPI_ISL_620910|2020 .................... .................... .................... .................... .................... ....................
P 2256 hCoV-19/Denmark/DCGC-5586/2020|EPI_ISL_620911|2020 .................... .................... .................... .................... .................... ....................
P 2257 hCoV-19/Denmark/DCGC-5587/2020|EPI_ISL_620912|2020 .................... .................... .................... .................... .................... ....................
P 2258 hCoV-19/Denmark/DCGC-5589/2020|EPI_ISL_620914|2020 .................... .................... .................... .................... .................... ....................
P 2259 hCoV-19/Denmark/DCGC-5590/2020|EPI_ISL_620915|2020 .................... .................... .................... .................... .................... ....................
P 2260 hCoV-19/Denmark/DCGC-5591/2020|EPI_ISL_620916|2020 .................... .................... .................... .................... .................... ....................
P 2261 hCoV-19/Denmark/DCGC-5592/2020|EPI_ISL_620917|2020 .................... .................... .................... .................... .................... ....................
P 2262 hCoV-19/Denmark/DCGC-5593/2020|EPI_ISL_620918|2020 .................... .................... .................... .................... .................... ....................
P 2263 hCoV-19/Denmark/DCGC-5594/2020|EPI_ISL_620919|2020 .................... .................... .................... .................... .................... ....................
P 2264 hCoV-19/Denmark/DCGC-5595/2020|EPI_ISL_620920|2020 .................... .................... .................... .................... .................... ....................
P 2265 hCoV-19/Denmark/DCGC-5596/2020|EPI_ISL_620921|2020 .................... .................... .................... .................... .................... ....................
P 2266 hCoV-19/Denmark/DCGC-5597/2020|EPI_ISL_620922|2020 .................... .................... .................... .................... .................... ....................
P 2267 hCoV-19/Denmark/DCGC-5599/2020|EPI_ISL_620924|2020 .................... .................... .................... .................... .................... ....................
P 2268 hCoV-19/Denmark/DCGC-5600/2020|EPI_ISL_620925|2020 .................... .................... .................... .................... .................... ....................
P 2269 hCoV-19/Denmark/DCGC-5601/2020|EPI_ISL_620926|2020 .................... .................... .................... .................... .................... ....................
P 2270 hCoV-19/Denmark/DCGC-5602/2020|EPI_ISL_620927|2020 .................... .................... .................... .................... .................... ....................
P 2271 hCoV-19/Denmark/DCGC-5603/2020|EPI_ISL_620928|2020 .................... .................... .................... .................... .T.................. ....................
P 2272 hCoV-19/Denmark/DCGC-5604/2020|EPI_ISL_620929|2020 .................... .................... .................... .................... .................... ....................
P 2273 hCoV-19/Denmark/DCGC-5605/2020|EPI_ISL_620930|2020 .................... .................... .................... .................... .................... ....................
P 2274 hCoV-19/Denmark/DCGC-5607/2020|EPI_ISL_620932|2020 .................... .................... .................... .................... .................... ....................
P 2275 hCoV-19/Denmark/DCGC-5608/2020|EPI_ISL_620933|2020 .................... .................... .................... .................... .................... ....................
```

| | | | | | | | | |
|---|---|---|---|---|---|---|---|---|
| P | 2276 | hCoV-19/Denmark/DCGC-5609/2020\|EPI_ISL_620934\|2020 | ···················· | ···················· | ····················· | ···················· | ···················· | ···················· |
| P | 2277 | hCoV-19/Denmark/DCGC-5610/2020\|EPI_ISL_620935\|2020 | ···················· | ···················· | ····················· | ···················· | ···················· | ···················· |
| P | 2278 | hCoV-19/Denmark/DCGC-5611/2020\|EPI_ISL_620936\|2020 | ···················· | ···················· | ····················· | ···················· | ···················· | ···················· |
| P | 2279 | hCoV-19/Denmark/DCGC-5612/2020\|EPI_ISL_620937\|2020 | ···················· | ···················· | ····················· | ···················· | ···················· | ···················· |
| P | 2280 | hCoV-19/Denmark/DCGC-5613/2020\|EPI_ISL_620938\|2020 | ···················· | ···················· | ····················· | ···················· | ···················· | ···················· |
| P | 2281 | hCoV-19/Denmark/DCGC-5614/2020\|EPI_ISL_620939\|2020 | ···················· | ···················· | ····················· | ···················· | ···················· | ···················· |
| P | 2282 | hCoV-19/Denmark/DCGC-5615/2020\|EPI_ISL_620940\|2020 | ···················· | ···················· | ····················· | ···················· | ···················· | ···················· |
| P | 2283 | hCoV-19/Denmark/DCGC-5616/2020\|EPI_ISL_620941\|2020 | ···················· | ···················· | ····················· | ···················· | ···················· | ···················· |
| P | 2284 | hCoV-19/Denmark/DCGC-5617/2020\|EPI_ISL_620942\|2020 | ···················· | ···················· | ····················· | ···················· | ···················· | ···················· |
| P | 2285 | hCoV-19/Denmark/DCGC-5618/2020\|EPI_ISL_620943\|2020 | ···················· | ···················· | ····················· | ···················· | ···················· | ···················· |
| P | 2286 | hCoV-19/Denmark/DCGC-5619/2020\|EPI_ISL_620944\|2020 | ···················· | ···················· | ····················· | ···················· | ···················· | ···················· |
| P | 2287 | hCoV-19/Denmark/DCGC-5620/2020\|EPI_ISL_620945\|2020 | ···················· | ···················· | ····················· | ···················· | ···················· | ···················· |
| P | 2288 | hCoV-19/Denmark/DCGC-5621/2020\|EPI_ISL_620946\|2020 | ···················· | ···················· | ····················· | ···················· | ···················· | ···················· |
| P | 2289 | hCoV-19/Denmark/DCGC-5622/2020\|EPI_ISL_620947\|2020 | ···················· | ···················· | ····················· | ···················· | ···················· | ···················· |
| P | 2290 | hCoV-19/Denmark/DCGC-5623/2020\|EPI_ISL_620948\|2020 | ···················· | ···················· | ····················· | ···················· | ···················· | ···················· |
| P | 2291 | hCoV-19/Denmark/DCGC-5624/2020\|EPI_ISL_620949\|2020 | ···················· | ···················· | ····················· | ···················· | ···················· | ···················· |
| P | 2292 | hCoV-19/Denmark/DCGC-5625/2020\|EPI_ISL_620950\|2020 | ···················· | ···················· | ····················· | ···················· | ···················· | ···················· |
| P | 2293 | hCoV-19/Denmark/DCGC-5626/2020\|EPI_ISL_620951\|2020 | ···················· | ···················· | ····················· | ···················· | ···················· | ···················· |
| P | 2294 | hCoV-19/Denmark/DCGC-5794/2020\|EPI_ISL_620981\|2020 | ···················· | ···················· | ····················· | ···················· | ···················· | ···················· |
| P | 2295 | hCoV-19/Denmark/DCGC-5796/2020\|EPI_ISL_620983\|2020 | ···················· | ···················· | ····················· | ···················· | ···················· | ···················· |
| P | 2296 | hCoV-19/Denmark/DCGC-5797/2020\|EPI_ISL_620984\|2020 | ···················· | ···················· | ····················· | ···················· | ···················· | ···················· |
| P | 2297 | hCoV-19/USA/UT-UPHL-201130447/2020\|EPI_ISL_623190\| | ···················· | ···················· | ····················· | ···················· | ···················· | ···················· |
| P | 2298 | hCoV-19/USA/UT-UPHL-201123076/2020\|EPI_ISL_623191\| | ···················· | ···················· | ····················· | ···················· | ···················· | ···················· |
| P | 2299 | hCoV-19/USA/UT-UPHL-201111436/2020\|EPI_ISL_623192\| | ···················· | ···················· | ····················· | ···················· | ···················· | ···················· |
| P | 2300 | hCoV-19/USA/UT-UPHL-201105159/2020\|EPI_ISL_623193\| | ···················· | ···················· | ····················· | ···················· | ···················· | ···················· |
| P | 2301 | hCoV-19/USA/UT-UPHL-201117690/2020\|EPI_ISL_623195\| | ···················· | ···················· | ····················· | ···················· | ···················· | ···················· |
| P | 2302 | hCoV-19/USA/UT-UPHL-201125896/2020\|EPI_ISL_623196\| | ···················· | ···················· | ····················· | ···················· | ···················· | ···················· |
| P | 2303 | hCoV-19/USA/UT-UPHL-201104021/2020\|EPI_ISL_623197\| | ···················· | ···················· | ····················· | ···················· | ···················· | ···················· |
| P | 2304 | hCoV-19/USA/UT-UPHL-201110657/2020\|EPI_ISL_623199\| | ···················· | ···················· | ····················· | ···················· | ···················· | ···················· |
| P | 2305 | hCoV-19/Denmark/DCGC-4402/2020\|EPI_ISL_615400\|2020 | ···················· | ···················· | ····················· | ···················· | ···················· | ···················· |
| P | 2306 | hCoV-19/Denmark/DCGC-4403/2020\|EPI_ISL_615401\|2020 | ···················· | ···················· | ····················· | ···················· | ···················· | ···················· |
| P | 2307 | hCoV-19/Denmark/DCGC-4404/2020\|EPI_ISL_615402\|2020 | ···················· | ···················· | ····················· | ···················· | ···················· | ···················· |
| P | 2308 | hCoV-19/Denmark/DCGC-4405/2020\|EPI_ISL_615403\|2020 | ···················· | ···················· | ····················· | ···················· | ···················· | ···················· |
| P | 2309 | hCoV-19/Denmark/DCGC-1717/2020\|EPI_ISL_615404\|2020 | ···················· | ···················· | ····················· | ···················· | ···················· | ···················· |
| P | 2310 | hCoV-19/Denmark/DCGC-1743/2020\|EPI_ISL_615405\|2020 | ···················· | ···················· | ····················· | ···················· | ···················· | ···················· |
| P | 2311 | hCoV-19/Denmark/DCGC-3635/2020\|EPI_ISL_615406\|2020 | ···················· | ···················· | ····················· | ···················· | ···················· | ···················· |
| P | 2312 | hCoV-19/Denmark/DCGC-3636/2020\|EPI_ISL_615407\|2020 | ···················· | ···················· | ····················· | ···················· | ···················· | ···················· |
| P | 2313 | hCoV-19/Denmark/DCGC-3637/2020\|EPI_ISL_615408\|2020 | ···················· | ···················· | ····················· | ···················· | ···················· | ···················· |
| P | 2314 | hCoV-19/Denmark/DCGC-3638/2020\|EPI_ISL_615409\|2020 | ···················· | ···················· | ····················· | ···················· | ···················· | ···················· |
| P | 2315 | hCoV-19/Denmark/DCGC-3639/2020\|EPI_ISL_615410\|2020 | ···················· | ···················· | ····················· | ···················· | ···················· | ···················· |
| P | 2316 | hCoV-19/Denmark/DCGC-3640/2020\|EPI_ISL_615411\|2020 | ···················· | ···················· | ····················· | ···················· | ···················· | ···················· |
| P | 2317 | hCoV-19/Denmark/DCGC-3641/2020\|EPI_ISL_615412\|2020 | ···················· | ···················· | ····················· | ···················· | ···················· | ···················· |
| P | 2318 | hCoV-19/Denmark/DCGC-3642/2020\|EPI_ISL_615413\|2020 | ···················· | ···················· | ····················· | ···················· | ···················· | ···················· |
| P | 2319 | hCoV-19/Denmark/DCGC-3643/2020\|EPI_ISL_615414\|2020 | ···················· | ···················· | ····················· | ···················· | ···················· | ···················· |
| P | 2320 | hCoV-19/Denmark/DCGC-6303/2020\|EPI_ISL_615415\|2020 | ···················· | ···················· | ····················· | ···················· | ···················· | ···················· |
| P | 2321 | hCoV-19/Denmark/DCGC-3644/2020\|EPI_ISL_615416\|2020 | ···················· | ···················· | ····················· | ···················· | ···················· | ···················· |
| P | 2322 | hCoV-19/Denmark/DCGC-3645/2020\|EPI_ISL_615417\|2020 | ···················· | ···················· | ····················· | ···················· | ···················· | ···················· |
| P | 2323 | hCoV-19/Denmark/DCGC-3646/2020\|EPI_ISL_615418\|2020 | ···················· | ·A·················· | ····················· | ···················· | ···················· | ··A················· |
| P | 2324 | hCoV-19/Denmark/DCGC-3647/2020\|EPI_ISL_615419\|2020 | ···················· | ···················· | ····················· | ···················· | ···················· | ···················· |
| P | 2325 | hCoV-19/Denmark/DCGC-3648/2020\|EPI_ISL_615420\|2020 | ···················· | ·A·················· | ····················· | ···················· | ···················· | ··A················· |
| P | 2326 | hCoV-19/Denmark/DCGC-3649/2020\|EPI_ISL_615421\|2020 | ···················· | ···················· | ····················· | ···················· | ···················· | ···················· |
| P | 2327 | hCoV-19/Denmark/DCGC-3650/2020\|EPI_ISL_615422\|2020 | ···················· | ···················· | ····················· | ···················· | ···················· | ···················· |
| P | 2328 | hCoV-19/Denmark/DCGC-3651/2020\|EPI_ISL_615423\|2020 | ···················· | ···················· | ····················· | ···················· | ···················· | ···················· |
| P | 2329 | hCoV-19/Denmark/DCGC-3652/2020\|EPI_ISL_615424\|2020 | ···················· | ···················· | ····················· | ···················· | ···················· | ···················· |
| P | 2330 | hCoV-19/Denmark/DCGC-3653/2020\|EPI_ISL_615425\|2020 | ···················· | ···················· | ····················· | ···················· | ···················· | ···················· |
| P | 2331 | hCoV-19/Denmark/DCGC-3654/2020\|EPI_ISL_615426\|2020 | ···················· | ···················· | ····················· | ···················· | ···················· | ···················· |
| P | 2332 | hCoV-19/Denmark/DCGC-3655/2020\|EPI_ISL_615427\|2020 | ···················· | ···················· | ····················· | ···················· | ···················· | ···················· |
| P | 2333 | hCoV-19/Denmark/DCGC-3656/2020\|EPI_ISL_615428\|2020 | ···················· | ···················· | ····················· | ···················· | ···················· | ···················· |
| P | 2334 | hCoV-19/Denmark/DCGC-3657/2020\|EPI_ISL_615429\|2020 | ···················· | ···················· | ····················· | ···················· | ···················· | ···················· |
| P | 2335 | hCoV-19/Denmark/DCGC-3658/2020\|EPI_ISL_615430\|2020 | ···················· | ···················· | ····················· | ···················· | ···················· | ···················· |
| P | 2336 | hCoV-19/Denmark/DCGC-3659/2020\|EPI_ISL_615431\|2020 | ···················· | ···················· | ····················· | ···················· | ···················· | ···················· |
| P | 2337 | hCoV-19/Denmark/DCGC-3660/2020\|EPI_ISL_615432\|2020 | ···················· | ···················· | ····················· | ···················· | ···················· | ···················· |
| P | 2338 | hCoV-19/Denmark/DCGC-3661/2020\|EPI_ISL_615433\|2020 | ···················· | ···················· | ····················· | ···················· | ···················· | ···················· |
| P | 2339 | hCoV-19/Denmark/DCGC-3662/2020\|EPI_ISL_615434\|2020 | ···················· | ···················· | ····················· | ···················· | ···················· | ···················· |
| P | 2340 | hCoV-19/Denmark/DCGC-3663/2020\|EPI_ISL_615435\|2020 | ···················· | ···················· | ····················· | ···················· | ···················· | ···················· |
| P | 2341 | hCoV-19/Denmark/DCGC-6304/2020\|EPI_ISL_615437\|2020 | ···················· | ···················· | ····················· | ···················· | ···················· | ···················· |
| P | 2342 | hCoV-19/Denmark/DCGC-3666/2020\|EPI_ISL_615439\|2020 | ···················· | ···················· | ····················· | ···················· | ···················· | ···················· |
| P | 2343 | hCoV-19/Denmark/DCGC-3667/2020\|EPI_ISL_615440\|2020 | ···················· | ···················· | ····················· | ···················· | ···················· | ···················· |
| P | 2344 | hCoV-19/Denmark/DCGC-3668/2020\|EPI_ISL_615441\|2020 | ···················· | ···················· | ····················· | ···················· | ···················· | ···················· |
| P | 2345 | hCoV-19/Denmark/DCGC-3669/2020\|EPI_ISL_615442\|2020 | ···················· | ···················· | ····················· | ···················· | ···················· | ···················· |
| P | 2346 | hCoV-19/Denmark/DCGC-3670/2020\|EPI_ISL_615443\|2020 | ···················· | ···················· | ····················· | ···················· | ···················· | ···················· |
| P | 2347 | hCoV-19/Denmark/DCGC-3671/2020\|EPI_ISL_615444\|2020 | ···················· | ···················· | ····················· | ···················· | ···················· | ···················· |
| P | 2348 | hCoV-19/Denmark/DCGC-3672/2020\|EPI_ISL_615445\|2020 | ···················· | ···················· | ····················· | ···················· | ···················· | ···················· |
| P | 2349 | hCoV-19/Denmark/DCGC-3673/2020\|EPI_ISL_615446\|2020 | ···················· | ···················· | ····················· | ···················· | ···················· | ···················· |
| P | 2350 | hCoV-19/Denmark/DCGC-3674/2020\|EPI_ISL_615447\|2020 | ···················· | ···················· | ····················· | ···················· | ···················· | ···················· |
| P | 2351 | hCoV-19/Denmark/DCGC-3677/2020\|EPI_ISL_615450\|2020 | ···················· | ···················· | ····················· | ···················· | ···················· | ···················· |
| P | 2352 | hCoV-19/Denmark/DCGC-3679/2020\|EPI_ISL_615454\|2020 | ···················· | ···················· | ····················· | ···················· | ···················· | ···················· |
| P | 2353 | hCoV-19/Denmark/DCGC-3680/2020\|EPI_ISL_615455\|2020 | ···················· | ···················· | ····················· | ···················· | ···················· | ···················· |
| P | 2354 | hCoV-19/Denmark/DCGC-3682/2020\|EPI_ISL_615457\|2020 | ···················· | ···················· | ····················· | ···················· | ···················· | ···················· |
| P | 2355 | hCoV-19/Denmark/DCGC-3683/2020\|EPI_ISL_615458\|2020 | ···················· | ···················· | ····················· | ···················· | ···················· | ···················· |

| | | | | | | | | | |
|---|---|---|---|---|---|---|---|---|---|
| P | 2356 | hCoV-19/Denmark/DCGC-3684/2020\|EPI_ISL_615459\|2020 | ···················· | ···················· | ···················· | ···················· | ···················· | ···················· | |
| P | 2357 | hCoV-19/Denmark/DCGC-3712/2020\|EPI_ISL_615490\|2020 | ···················· | ···················· | ···················· | ···················· | ···················· | ···················· | |
| P | 2358 | hCoV-19/Denmark/DCGC-3713/2020\|EPI_ISL_615491\|2020 | ···················· | ···················· | ···················· | ···················· | ···················· | ···················· | |
| P | 2359 | hCoV-19/Denmark/DCGC-3714/2020\|EPI_ISL_615492\|2020 | ···················· | ···················· | ···················· | ···················· | ···················· | ···················· | |
| P | 2360 | hCoV-19/Denmark/DCGC-3715/2020\|EPI_ISL_615493\|2020 | ···················· | ···················· | ···················· | ···················· | ···················· | ···················· | |
| P | 2361 | hCoV-19/Denmark/DCGC-3716/2020\|EPI_ISL_615494\|2020 | ···················· | ···················· | ···················· | ···················· | ···················· | ···················· | |
| P | 2362 | hCoV-19/Denmark/DCGC-3717/2020\|EPI_ISL_615495\|2020 | ···················· | ···················· | ···················· | ···················· | ···················· | ···················· | |
| P | 2363 | hCoV-19/Denmark/DCGC-3718/2020\|EPI_ISL_615496\|2020 | ···················· | ···················· | ···················· | ···················· | ···················· | ···················· | |
| P | 2364 | hCoV-19/Denmark/DCGC-3719/2020\|EPI_ISL_615497\|2020 | ···················· | ···················· | ···················· | ···················· | ···················· | ···················· | |
| P | 2365 | hCoV-19/Denmark/DCGC-3720/2020\|EPI_ISL_615498\|2020 | ···················· | ···················· | ···················· | ···················· | ···················· | ···················· | |
| P | 2366 | hCoV-19/Denmark/DCGC-3721/2020\|EPI_ISL_615499\|2020 | ···················· | ···················· | ···················· | ···················· | ···················· | ···················· | |
| P | 2367 | hCoV-19/Denmark/DCGC-2939/2020\|EPI_ISL_616700\|2020 | ···················· | ···················· | ···················· | ···················· | ···················· | ···················· | |
| P | 2368 | hCoV-19/Denmark/DCGC-2940/2020\|EPI_ISL_616701\|2020 | ···················· | ···················· | ···················· | ···················· | ···················· | ···················· | |
| P | 2369 | hCoV-19/Denmark/DCGC-2942/2020\|EPI_ISL_616703\|2020 | ···················· | ···················· | ···················· | ···················· | ···················· | ···················· | |
| P | 2370 | hCoV-19/Denmark/DCGC-2943/2020\|EPI_ISL_616704\|2020 | ···················· | ···················· | ···················· | ···················· | ···················· | ···················· | |
| P | 2371 | hCoV-19/Denmark/DCGC-2944/2020\|EPI_ISL_616705\|2020 | ···················· | ···················· | ···················· | ···················· | ···················· | ···················· | |
| P | 2372 | hCoV-19/Denmark/DCGC-2945/2020\|EPI_ISL_616706\|2020 | ···················· | ···················· | ···················· | ···················· | ···················· | ···················· | |
| P | 2373 | hCoV-19/Denmark/DCGC-2946/2020\|EPI_ISL_616707\|2020 | ···················· | ···················· | ···················· | ···················· | ···················· | ···················· | |
| P | 2374 | hCoV-19/Denmark/DCGC-2947/2020\|EPI_ISL_616708\|2020 | ···················· | ···················· | ···················· | ···················· | ···················· | ···················· | |
| P | 2375 | hCoV-19/Denmark/DCGC-2948/2020\|EPI_ISL_616709\|2020 | ···················· | ···················· | ···················· | ···················· | ···················· | ···················· | |
| P | 2376 | hCoV-19/Denmark/DCGC-2949/2020\|EPI_ISL_616710\|2020 | ···················· | ···················· | ···················· | ···················· | ···················· | ···················· | |
| P | 2377 | hCoV-19/Denmark/DCGC-2950/2020\|EPI_ISL_616711\|2020 | ···················· | ···················· | ···················· | ···················· | ···················· | ···················· | |
| P | 2378 | hCoV-19/Denmark/DCGC-2951/2020\|EPI_ISL_616712\|2020 | ···················· | ···················· | ···················· | ···················· | ···················· | ···················· | |
| P | 2379 | hCoV-19/Denmark/DCGC-2952/2020\|EPI_ISL_616713\|2020 | ···················· | ···················· | ···················· | ···················· | ···················· | ···················· | |
| P | 2380 | hCoV-19/Denmark/DCGC-2954/2020\|EPI_ISL_616715\|2020 | ···················· | ···················· | ···················· | ···················· | ···················· | ···················· | |
| P | 2381 | hCoV-19/Denmark/DCGC-2955/2020\|EPI_ISL_616716\|2020 | ···················· | ···················· | ···················· | ···················· | ···················· | ···················· | |
| P | 2382 | hCoV-19/Denmark/DCGC-2956/2020\|EPI_ISL_616717\|2020 | ···················· | ···················· | ···················· | ···················· | ···················· | ···················· | |
| P | 2383 | hCoV-19/Denmark/DCGC-2930/2020\|EPI_ISL_616718\|2020 | ···················· | ···················· | ···················· | ···················· | ···················· | ···················· | |
| P | 2384 | hCoV-19/Denmark/DCGC-2957/2020\|EPI_ISL_616719\|2020 | ···················· | ·A·················· | ···················· | ···················· | ···················· | ··A················· | |
| P | 2385 | hCoV-19/Denmark/DCGC-2958/2020\|EPI_ISL_616720\|2020 | ···················· | ···················· | ···················· | ···················· | ···················· | ···················· | |
| P | 2386 | hCoV-19/Denmark/DCGC-2959/2020\|EPI_ISL_616721\|2020 | ···················· | ···················· | ···················· | ···················· | ···················· | ···················· | |
| P | 2387 | hCoV-19/Denmark/DCGC-2960/2020\|EPI_ISL_616722\|2020 | ···················· | ···················· | ···················· | ···················· | ···················· | ···················· | |
| P | 2388 | hCoV-19/Denmark/DCGC-2961/2020\|EPI_ISL_616723\|2020 | ···················· | ···················· | ···················· | ···················· | ···················· | ···················· | |
| P | 2389 | hCoV-19/Denmark/DCGC-2963/2020\|EPI_ISL_616725\|2020 | ···················· | ···················· | ···················· | ···················· | ···················· | ···················· | |
| P | 2390 | hCoV-19/Denmark/DCGC-2964/2020\|EPI_ISL_616726\|2020 | ···················A | ···················· | ···················· | ···················· | ···················· | ···················· | 1 |
| P | 2391 | hCoV-19/Denmark/DCGC-2966/2020\|EPI_ISL_616728\|2020 | ···················· | ···················· | ···················· | ···················· | ···················· | ···················· | |
| P | 2392 | hCoV-19/Denmark/DCGC-2967/2020\|EPI_ISL_616729\|2020 | ···················· | ···················· | ···················· | ···················· | ···················· | ···················· | |
| P | 2393 | hCoV-19/Denmark/DCGC-2969/2020\|EPI_ISL_616731\|2020 | ···················· | ···················· | ···················· | ···················· | ···················· | ···················· | |
| P | 2394 | hCoV-19/Denmark/DCGC-2970/2020\|EPI_ISL_616732\|2020 | ···················· | ···················· | ···················· | ···················· | ···················· | ···················· | |
| P | 2395 | hCoV-19/Denmark/DCGC-2971/2020\|EPI_ISL_616733\|2020 | ···················· | ···················· | ···················· | ···················· | ···················· | ···················· | |
| P | 2396 | hCoV-19/Denmark/DCGC-2972/2020\|EPI_ISL_616734\|2020 | ···················· | ···················· | ···················· | ···················· | ···················· | ···················· | |
| P | 2397 | hCoV-19/Denmark/DCGC-2973/2020\|EPI_ISL_616735\|2020 | ···················· | ···················· | ···················· | ···················· | ···················· | ···················· | |
| P | 2398 | hCoV-19/Denmark/DCGC-2974/2020\|EPI_ISL_616736\|2020 | ···················· | ···················· | ···················· | ···················· | ···················· | ···················· | |
| P | 2399 | hCoV-19/Denmark/DCGC-2975/2020\|EPI_ISL_616737\|2020 | ···················· | ···················· | ···················· | ···················· | ···················· | ···················· | |
| P | 2400 | hCoV-19/Denmark/DCGC-2976/2020\|EPI_ISL_616738\|2020 | ···················· | ···················· | ···················· | ···················· | ···················· | ···················· | |
| P | 2401 | hCoV-19/Denmark/DCGC-2977/2020\|EPI_ISL_616739\|2020 | ···················· | ···················· | ···················· | ···················· | ···················· | ···················· | |
| P | 2402 | hCoV-19/Denmark/DCGC-2978/2020\|EPI_ISL_616740\|2020 | ···················· | ···················· | ···················· | ···················· | ···················· | ···················· | |
| P | 2403 | hCoV-19/Denmark/DCGC-2980/2020\|EPI_ISL_616742\|2020 | ···················· | ···················· | ···················· | ···················· | ···················· | ···················· | |
| P | 2404 | hCoV-19/Denmark/DCGC-2981/2020\|EPI_ISL_616743\|2020 | ···················· | ·A·················· | ···················· | ···················· | ···················· | ··A················· | |
| P | 2405 | hCoV-19/Denmark/DCGC-2982/2020\|EPI_ISL_616744\|2020 | ···················· | ·A·················· | ···················· | ···················· | ···················· | ··A················· | |
| P | 2406 | hCoV-19/Denmark/DCGC-2985/2020\|EPI_ISL_616747\|2020 | ···················· | ···················· | ···················· | ···················· | ···················· | ···················· | |
| P | 2407 | hCoV-19/Denmark/DCGC-2931/2020\|EPI_ISL_616750\|2020 | ···················· | ·A·················· | ···················· | ···················· | ···················· | ··A················· | |
| P | 2408 | hCoV-19/Denmark/DCGC-2932/2020\|EPI_ISL_616752\|2020 | ···················· | ···················· | ···················· | ···················· | ···················· | ···················· | |
| P | 2409 | hCoV-19/Denmark/DCGC-2989/2020\|EPI_ISL_616753\|2020 | ···················· | ···················· | ···················· | ···················· | ···················· | ···················· | |
| P | 2410 | hCoV-19/Denmark/DCGC-2990/2020\|EPI_ISL_616754\|2020 | ···················· | ···················· | ···················· | ···················· | ···················· | ···················· | |
| P | 2411 | hCoV-19/Denmark/DCGC-2991/2020\|EPI_ISL_616755\|2020 | ···················· | ···················· | ···················· | ···················· | ···················· | ···················· | |
| P | 2412 | hCoV-19/Denmark/DCGC-2992/2020\|EPI_ISL_616756\|2020 | ···················· | ···················· | ···················· | ···················· | ···················· | ···················· | |
| P | 2413 | hCoV-19/Denmark/DCGC-2994/2020\|EPI_ISL_616758\|2020 | ···················· | ···················· | ···················· | ···················· | ···················· | ···················· | |
| P | 2414 | hCoV-19/Denmark/DCGC-2995/2020\|EPI_ISL_616759\|2020 | ···················· | ···················· | ···················· | ···················· | ···················· | ···················· | |
| P | 2415 | hCoV-19/Denmark/DCGC-2999/2020\|EPI_ISL_616763\|2020 | ···················· | ···················· | ···················· | ···················· | ···················· | ···················· | |
| P | 2416 | hCoV-19/Denmark/DCGC-3208/2020\|EPI_ISL_616764\|2020 | ···················· | ···················· | ···················· | ···················· | ···················· | ···················· | |
| P | 2417 | hCoV-19/Denmark/DCGC-3209/2020\|EPI_ISL_616765\|2020 | ···················· | ···················· | ···················· | ···················· | ···················· | ···················· | |
| P | 2418 | hCoV-19/Denmark/DCGC-3210/2020\|EPI_ISL_616766\|2020 | ···················· | ···················· | ···················· | ···················· | ···················· | ···················· | |
| P | 2419 | hCoV-19/Denmark/DCGC-3211/2020\|EPI_ISL_616767\|2020 | ···················· | ···················· | ···················· | ···················· | ···················· | ···················· | |
| P | 2420 | hCoV-19/Denmark/DCGC-3212/2020\|EPI_ISL_616768\|2020 | ···················· | ···················· | ···················· | ···················· | ···················· | ···················· | |
| P | 2421 | hCoV-19/Denmark/DCGC-3213/2020\|EPI_ISL_616769\|2020 | ···················· | ·A·················· | ···················· | ···················· | ···················· | ···················· | |
| P | 2422 | hCoV-19/Denmark/DCGC-3214/2020\|EPI_ISL_616770\|2020 | ···················· | ···················· | ···················· | ···················· | ···················· | ···················· | |
| P | 2423 | hCoV-19/Denmark/DCGC-3216/2020\|EPI_ISL_616772\|2020 | ········T··········· | ···················· | ···················· | ···················· | ···················· | ···················· | |
| P | 2424 | hCoV-19/Denmark/DCGC-3217/2020\|EPI_ISL_616773\|2020 | ···················· | ·A·················· | ···················· | ···················· | ···················· | ···················· | |
| P | 2425 | hCoV-19/Denmark/DCGC-3280/2020\|EPI_ISL_616774\|2020 | ···················· | ···················· | ···················· | ···················· | ···················· | ···················· | |
| P | 2426 | hCoV-19/Denmark/DCGC-3001/2020\|EPI_ISL_616777\|2020 | ···················· | ·A·················· | ···················· | ···················· | ···················· | ··A················· | |
| P | 2427 | hCoV-19/Denmark/DCGC-3003/2020\|EPI_ISL_616779\|2020 | ···················· | ···················· | ···················· | ···················· | ···················· | ···················· | |
| P | 2428 | hCoV-19/Denmark/DCGC-3005/2020\|EPI_ISL_616781\|2020 | ···················· | ···················· | ···················· | ···················· | ···················· | ···················· | |
| P | 2429 | hCoV-19/Denmark/DCGC-3008/2020\|EPI_ISL_616784\|2020 | ···················· | ···················· | ···················· | ···················· | ···················· | ···················· | |
| P | 2430 | hCoV-19/Denmark/DCGC-3009/2020\|EPI_ISL_616785\|2020 | ···················· | ·A·················· | ···················· | ···················· | ···················· | ···················· | |
| P | 2431 | hCoV-19/Denmark/DCGC-3010/2020\|EPI_ISL_616786\|2020 | ···················· | ···················· | ···················· | ···················· | ···················· | ···················· | |
| P | 2432 | hCoV-19/Denmark/DCGC-3240/2020\|EPI_ISL_616787\|2020 | ···················· | ···················· | ···················· | ···················· | ···················· | ···················· | |
| P | 2433 | hCoV-19/Denmark/DCGC-3241/2020\|EPI_ISL_616789\|2020 | ···················· | ···················· | ···················· | ···················· | ···················· | ···················· | |
| P | 2434 | hCoV-19/Denmark/DCGC-8232/2020\|EPI_ISL_617260\|2020 | ···················· | ···················· | ···················· | ···················· | ···················· | ···················· | |
| P | 2435 | hCoV-19/Denmark/DCGC-8215/2020\|EPI_ISL_617261\|2020 | ···················· | ···················· | ···················· | ···················· | ···················· | ···················· | |

P 2436 hCoV-19/Denmark/DCGC-8193/2020|EPI_ISL_617264|2020 ···················· ···················· ····················· ···················· ······T············· ····················
P 2437 hCoV-19/Denmark/DCGC-8201/2020|EPI_ISL_617265|2020 ···················· ···················· ····················· ···················· ···················· ····················
P 2438 hCoV-19/Denmark/DCGC-7707/2020|EPI_ISL_618590|2020 ···················· ···················· ····················· ···················· ···················· ····················
P 2439 hCoV-19/Denmark/DCGC-7712/2020|EPI_ISL_618591|2020 ···················· ···················· ····················· ···················· ···················· ····················
P 2440 hCoV-19/Denmark/DCGC-7719/2020|EPI_ISL_618592|2020 ···················· ···················· ····················· ···················· ···················· ····················
P 2441 hCoV-19/Denmark/DCGC-1580/2020|EPI_ISL_618595|2020 ···················· ···················· ····················· ···················· ···················· ····················
P 2442 hCoV-19/Denmark/DCGC-7678/2020|EPI_ISL_618596|2020 ···················· ···················· ····················· ···················· ···················· ····················
P 2444 hCoV-19/Denmark/DCGC-4623/2020|EPI_ISL_621292|2020 ···················· ···················· ····················· ···················· ···················· ····················
P 2445 hCoV-19/Denmark/DCGC-4624/2020|EPI_ISL_621293|2020 ···················· ···················· ····················· ···················· ···················· ····················
P 2446 hCoV-19/Denmark/DCGC-4625/2020|EPI_ISL_621294|2020 ···················· ···················· ····················· ···················· ···················· ····················
P 2447 hCoV-19/Denmark/DCGC-4626/2020|EPI_ISL_621295|2020 ···················· ···················· ····················· ···················· ···················· ····················
P 2448 hCoV-19/Denmark/DCGC-4627/2020|EPI_ISL_621296|2020 ···················· ···················· ····················· ···················· ···················· ····················
P 2449 hCoV-19/Denmark/DCGC-4628/2020|EPI_ISL_621297|2020 ···················· ···················· ····················· ···················· ···················· ····················
P 2450 hCoV-19/Denmark/DCGC-4629/2020|EPI_ISL_621298|2020 ···················· ···················· ····················· ···················· ···················· ····················
P 2451 hCoV-19/Denmark/DCGC-4630/2020|EPI_ISL_621299|2020 ···················· ···················· ····················· ···················· ···················· ····················
P 2452 hCoV-19/France/ARA-CDP82132/2020|EPI_ISL_623100|20 ···················· ···················· ····················· ···················· ···················· ····················
P 2453 hCoV-19/France/ARA-CDP82221/2020|EPI_ISL_623101|20 ···················· ···················· ····················· ···················· ···················· ····················
P 2454 hCoV-19/France/ARA-CDP82284/2020|EPI_ISL_623102|20 ···················· ···················· ····················· ···················· ···················· ····················
P 2455 hCoV-19/England/ALDP-AA3ADA/2020|EPI_ISL_624400|20 ···················· ···················· ····················· ···················· ···················· ····················
P 2456 hCoV-19/England/ALDP-AA1A09/2020|EPI_ISL_624401|20 ···················· ···················· ····················· ···················· ···················· ····················
P 2457 hCoV-19/England/ALDP-AA3A9E/2020|EPI_ISL_624402|20 ···················· ···················· ····················· ···················· ···················· ····················
P 2458 hCoV-19/England/ALDP-AA3CE3/2020|EPI_ISL_624403|20 ···················· ···················· ····················· ···················· ···················· ····················
P 2459 hCoV-19/England/ALDP-AA1EB2/2020|EPI_ISL_624404|20 ···················· ···················· ····················· ···················· ···················· ····················
P 2460 hCoV-19/England/ALDP-AA4383/2020|EPI_ISL_624405|20 ···················· ···················· ····················· ···················· ···················· ····················
P 2461 hCoV-19/England/ALDP-AA27B6/2020|EPI_ISL_624406|20 ···················· ···················· ····················· ···················· ···················· ····················
P 2462 hCoV-19/England/ALDP-AA474B/2020|EPI_ISL_624407|20 ···················· ···················· ····················· ···················· ···················· ····················
P 2463 hCoV-19/England/ALDP-AA1AEB/2020|EPI_ISL_624408|20 ···················· ···················· ····················· ···················· ···················· ····················
P 2464 hCoV-19/England/ALDP-AA3E38/2020|EPI_ISL_624409|20 ···················· ···················· ····················· ···················· ···················· ····················
P 2465 hCoV-19/England/ALDP-AA46D5/2020|EPI_ISL_624410|20 ···················· ···················· ····················· ···················· ···················· ····················
P 2466 hCoV-19/England/ALDP-AA3AF8/2020|EPI_ISL_624411|20 ···················· ·A·················· ····················· ···················· ···················· ····················
P 2467 hCoV-19/England/ALDP-AA1043/2020|EPI_ISL_624412|20 ···················· ···················· ····················· ···················· ···················· ····················
P 2468 hCoV-19/England/ALDP-AA3C3E/2020|EPI_ISL_624413|20 ···················· ···················· ····················· ···················· ···················· ····················
P 2469 hCoV-19/England/ALDP-AA10CB/2020|EPI_ISL_624414|20 ···················· ···················· ····················· ···················· ···················· ····················
P 2470 hCoV-19/England/ALDP-AA1061/2020|EPI_ISL_624415|20 ···················· ···················· ····················· ···················· ···················· ····················
P 2471 hCoV-19/England/ALDP-AA431A/2020|EPI_ISL_624416|20 ···················· ···················· ····················· ···················· ···················· ····················
P 2472 hCoV-19/England/ALDP-AA14FC/2020|EPI_ISL_624417|20 ···················· ···················· ····················· ···················· ···················· ····················
P 2473 hCoV-19/England/ALDP-AA43A1/2020|EPI_ISL_624419|20 ···················· ···················· ····················· ···················· ···················· ····················
P 2474 hCoV-19/England/ALDP-AA39A0/2020|EPI_ISL_624420|20 ···················· ·A·················· ····················· ···················· ···················· ···A················
P 2475 hCoV-19/England/ALDP-AA1FBF/2020|EPI_ISL_624421|20 ···················· ···················· ····················· ···················· ···················· ····················
P 2476 hCoV-19/England/ALDP-AA1D3D/2020|EPI_ISL_624422|20 ···················· ···················· ····················· ···················· ···················· ····················
P 2477 hCoV-19/England/ALDP-AA3A34/2020|EPI_ISL_624423|20 ···················· ·A·······A·········· ····················· ···················· ···················· ····················
P 2478 hCoV-19/England/ALDP-AA3DFF/2020|EPI_ISL_624424|20 ···················· ···················· ····················· ···················· ···················· ····················
P 2479 hCoV-19/England/ALDP-AA3BAA/2020|EPI_ISL_624425|20 ···················· ···················· ····················· ···················· ···················· ····················
P 2480 hCoV-19/England/ALDP-AA44BD/2020|EPI_ISL_624426|20 ···················· ···················· ····················· ···················· ···················· ····················
P 2481 hCoV-19/England/ALDP-AA4620/2020|EPI_ISL_624427|20 ···················· ···················· ····················· ···················· ···················· ····················
P 2482 hCoV-19/England/ALDP-AA232B/2020|EPI_ISL_624428|20 ···················· ·A·················· ····················· ···················· ···················· ···A················
P 2483 hCoV-19/England/ALDP-AA1FDD/2020|EPI_ISL_624429|20 ···················· ···················· ····················· ···················· ···················· ····················
P 2484 hCoV-19/England/ALDP-AA1EEF/2020|EPI_ISL_624430|20 ···················· ···················· ····················· ···················· ···················· ····················
P 2485 hCoV-19/Wales/ALDP-AA4936/2020|EPI_ISL_624431|2020 ···················· ···················· ····················· ···················· ···················· ····················
P 2486 hCoV-19/England/ALDP-AA3B40/2020|EPI_ISL_624432|20 ···················· ···················· ····················· ···················· ···················· ····················
P 2487 hCoV-19/England/ALDP-AA22E2/2020|EPI_ISL_624433|20 ···················· ···················· ····················· ···················· ···················· ····················
P 2488 hCoV-19/England/ALDP-AA3C7A/2020|EPI_ISL_624434|20 ···················· ···················· ····················· ···················· ···················· ····················
P 2489 hCoV-19/England/ALDP-AA2394/2020|EPI_ISL_624435|20 ···················· ···················· ····················· ···················· ···················· ····················
P 2490 hCoV-19/England/ALDP-AA1F55/2020|EPI_ISL_624436|20 ···················· ···················· ····················· ···················· ···················· ····················
P 2491 hCoV-19/England/ALDP-AA1B06/2020|EPI_ISL_624437|20 ···················· ···················· ····················· ···················· ···················· ····················
P 2492 hCoV-19/England/ALDP-AA4BA9/2020|EPI_ISL_624438|20 ···················· ····A··············· ····················· ···················· ···················· ····················
P 2493 hCoV-19/Denmark/DCGC-8603/2020|EPI_ISL_625703|2020 ···················· ···················· ····················· ···················· ···················· ····················
P 2494 hCoV-19/Denmark/DCGC-8668/2020|EPI_ISL_625716|2020 ···················· ·A·················· ····················· ···················· ···················· ···A················
P 2495 hCoV-19/Denmark/DCGC-8647/2020|EPI_ISL_625732|2020 ···················· ···················· ····················· ···················· ···················· ····················
P 2496 hCoV-19/Denmark/DCGC-8497/2020|EPI_ISL_625733|2020 ···················· ···················· ····················· ···················· ···················· ····················
P 2497 hCoV-19/Denmark/DCGC-8791/2020|EPI_ISL_625734|2020 ···················· ···················· ····················· ···················· ···················· ····················
P 2498 hCoV-19/Denmark/DCGC-8870/2020|EPI_ISL_625735|2020 ···················· ···················· ····················· ···················· ···················· ····················
P 2499 hCoV-19/Denmark/DCGC-8860/2020|EPI_ISL_625736|2020 ···················· ···················· ····················· ···················· ···················· ····················
P 2500 hCoV-19/Denmark/DCGC-8496/2020|EPI_ISL_625737|2020 ···················· ···················· ····················· ···················· ···················· ····················
P 2501 hCoV-19/Denmark/DCGC-8613/2020|EPI_ISL_625738|2020 ···················· ···················· ····················· ···················· ···················· ····················
P 2502 hCoV-19/Denmark/DCGC-8855/2020|EPI_ISL_625739|2020 ···················· ···················· ····················· ···················· ···················· ····················
P 2503 hCoV-19/Denmark/DCGC-8970/2020|EPI_ISL_625740|2020 ···················· ···················· ····················· ···················· ···················· ····················
P 2504 hCoV-19/Denmark/DCGC-8966/2020|EPI_ISL_625741|2020 ···················· ···················· ····················· ···················· ···················· ····················
P 2505 hCoV-19/Denmark/DCGC-8652/2020|EPI_ISL_625746|2020 ···················· ···················· ····················· ···················· ···················· ····················
P 2506 hCoV-19/Denmark/DCGC-8645/2020|EPI_ISL_625752|2020 ···················· ···················· ····················· ···················· ···················· ····················
P 2507 hCoV-19/Denmark/DCGC-9005/2020|EPI_ISL_625756|2020 ···················· ···················· ····················· ···················· ···················· ····················
P 2508 hCoV-19/Denmark/DCGC-8494/2020|EPI_ISL_625757|2020 ···················· ···················· ····················· ···················· ···················· ····················
P 2509 hCoV-19/Denmark/DCGC-8511/2020|EPI_ISL_625758|2020 ···················· ···················· ····················· ···················· ···················· ····················
P 2510 hCoV-19/Denmark/DCGC-8519/2020|EPI_ISL_625759|2020 ···················· ···················· ····················· ···················· ···················· ····················
P 2511 hCoV-19/Denmark/DCGC-8524/2020|EPI_ISL_625760|2020 ···················· ···················· ····················· ···················· ···················· ····················
P 2512 hCoV-19/Denmark/DCGC-8531/2020|EPI_ISL_625761|2020 ···················· ···················· ····················· ···················· ···················· ····················
P 2513 hCoV-19/Denmark/DCGC-8535/2020|EPI_ISL_625762|2020 ···················· ···················· ····················· ···················· ···················· ····················
P 2514 hCoV-19/Denmark/DCGC-8495/2020|EPI_ISL_625763|2020 ···················· ···················· ····················· ···················· ···················· ····················
P 2515 hCoV-19/Denmark/DCGC-8512/2020|EPI_ISL_625764|2020 ···················· ···················· ····················· ···················· ···················· ····················
P 2516 hCoV-19/Denmark/DCGC-8515/2020|EPI_ISL_625765|2020 ···················· ···················· ····················· ···················· ···················· ····················

| | | | | | | | | |
|---|---|---|---|---|---|---|---|---|
| P | 2517 | hCoV-19/Denmark/DCGC-8532/2020\|EPI_ISL_625766\|2020 | ···················· | ···················· | ···················· | ···················· | ···················· | ···················· |
| P | 2518 | hCoV-19/Denmark/DCGC-8521/2020\|EPI_ISL_625767\|2020 | ···················· | ···················· | ···················· | ···················· | ···················· | ···················· |
| P | 2519 | hCoV-19/Denmark/DCGC-8525/2020\|EPI_ISL_625768\|2020 | ···················· | ···················· | ···················· | ···················· | ···················· | ···················· |
| P | 2520 | hCoV-19/Denmark/DCGC-8537/2020\|EPI_ISL_625769\|2020 | ···················· | ···················· | ···················· | ···················· | ···················· | ···················· |
| P | 2521 | hCoV-19/Denmark/DCGC-8883/2020\|EPI_ISL_626140\|2020 | ···················· | ···················· | ···················· | ···················· | ···················· | ···················· |
| P | 2522 | hCoV-19/Denmark/DCGC-8902/2020\|EPI_ISL_626141\|2020 | ···················· | ···················· | ···················· | ···················· | ···················· | ···················· |
| P | 2523 | hCoV-19/Denmark/DCGC-8910/2020\|EPI_ISL_626142\|2020 | ···················· | ···················· | ···················· | ···················· | ···················· | ···················· |
| P | 2524 | hCoV-19/Denmark/DCGC-8932/2020\|EPI_ISL_626143\|2020 | ···················· | ·A·················· | ···················· | ···················· | ···················· | ···················· |
| P | 2525 | hCoV-19/Denmark/DCGC-8940/2020\|EPI_ISL_626144\|2020 | ···················· | ···················· | ···················· | ···················· | ···················· | ···················· |
| P | 2526 | hCoV-19/Denmark/DCGC-8946/2020\|EPI_ISL_626145\|2020 | ···················· | ···················· | ···················· | ···················· | ···················· | ···················· |
| P | 2527 | hCoV-19/Denmark/DCGC-8877/2020\|EPI_ISL_626146\|2020 | ···················· | ·A·················· | ···················· | ···················· | ···················· | ···················· |
| P | 2528 | hCoV-19/Denmark/DCGC-8884/2020\|EPI_ISL_626147\|2020 | ···················· | ···················· | ···················· | ···················· | ···················· | ···················· |
| P | 2529 | hCoV-19/Denmark/DCGC-8891/2020\|EPI_ISL_626148\|2020 | ···················· | ···················· | ···················· | ···················· | ···················· | ···················· |
| P | 2530 | hCoV-19/Denmark/DCGC-8897/2020\|EPI_ISL_626149\|2020 | ···················· | ·A·················· | ···················· | ···················· | ···················· | ···················· |
| P | 2531 | hCoV-19/Denmark/DCGC-8919/2020\|EPI_ISL_626150\|2020 | ···················· | ·A·················· | ···················· | ···················· | ···················· | ···················· |
| P | 2532 | hCoV-19/Denmark/DCGC-8933/2020\|EPI_ISL_626151\|2020 | ···················· | ···················· | ···················· | ···················· | ···················· | ···················· |
| P | 2533 | hCoV-19/Denmark/DCGC-8941/2020\|EPI_ISL_626152\|2020 | ···················· | ···················· | ···················· | ···················· | ···················· | ···················· |
| P | 2534 | hCoV-19/Denmark/DCGC-8904/2020\|EPI_ISL_626153\|2020 | ···················· | ···················· | ···················· | ···················· | ···················· | ···················· |
| P | 2535 | hCoV-19/Denmark/DCGC-8912/2020\|EPI_ISL_626154\|2020 | ···················· | ···················· | ···················· | ···················· | ···················· | ···················· |
| P | 2536 | hCoV-19/Denmark/DCGC-8925/2020\|EPI_ISL_626155\|2020 | ···················· | ···················· | ···················· | ···················· | ···················· | ···················· |
| P | 2537 | hCoV-19/Denmark/DCGC-8934/2020\|EPI_ISL_626156\|2020 | ···················· | ···················· | ···················· | ···················· | ···················· | ···················· |
| P | 2538 | hCoV-19/Denmark/DCGC-8878/2020\|EPI_ISL_626157\|2020 | ···················· | ···················· | ···················· | ···················· | ···················· | ···················· |
| P | 2539 | hCoV-19/Denmark/DCGC-8920/2020\|EPI_ISL_626158\|2020 | ···················· | ···················· | ···················· | ···················· | ···················· | ···················· |
| P | 2540 | hCoV-19/Denmark/DCGC-8928/2020\|EPI_ISL_626159\|2020 | ···················· | ···················· | ···················· | ···················· | ···················· | ···················· |
| P | 2541 | hCoV-19/Denmark/DCGC-8935/2020\|EPI_ISL_626160\|2020 | ···················· | ···················· | ···················· | ···················· | ···················· | ···················· |
| P | 2542 | hCoV-19/Denmark/DCGC-8943/2020\|EPI_ISL_626161\|2020 | ···················· | ···················· | ···················· | ···················· | ···················· | ···················· |
| P | 2543 | hCoV-19/Denmark/DCGC-8879/2020\|EPI_ISL_626162\|2020 | ···················· | ···················· | ···················· | ···················· | ···················· | ···················· |
| P | 2544 | hCoV-19/Denmark/DCGC-8887/2020\|EPI_ISL_626163\|2020 | ··········N········· | ···················· | ···················· | ···················· | ···················· | ···················· |
| P | 2545 | hCoV-19/Denmark/DCGC-8893/2020\|EPI_ISL_626164\|2020 | ···················· | ···················· | ···················· | ···················· | ···················· | ···················· |
| P | 2546 | hCoV-19/Denmark/DCGC-8914/2020\|EPI_ISL_626165\|2020 | ···················· | ···················· | ···················· | ···················· | ···················· | ···················· |
| P | 2547 | hCoV-19/Denmark/DCGC-8929/2020\|EPI_ISL_626166\|2020 | ···················· | ···················· | ···················· | ···················· | ···················· | ···················· |
| P | 2548 | hCoV-19/Denmark/DCGC-8936/2020\|EPI_ISL_626167\|2020 | ···················· | ···················· | ···················· | ···················· | ···················· | ···················· |
| P | 2549 | hCoV-19/Denmark/DCGC-8944/2020\|EPI_ISL_626168\|2020 | ···················· | ·A·················· | ···················· | ···················· | ···················· | ···················· |
| P | 2550 | hCoV-19/Denmark/DCGC-8977/2020\|EPI_ISL_626169\|2020 | ···················· | ···················· | ···················· | ···················· | ···················· | ···················· |
| P | 2551 | hCoV-19/Denmark/DCGC-8995/2020\|EPI_ISL_626170\|2020 | ···················· | ···················· | ···················· | ···················· | ···················· | ···················· |
| P | 2552 | hCoV-19/Denmark/DCGC-9006/2020\|EPI_ISL_626171\|2020 | ···················· | ···················· | ···················· | ···················· | ···················· | ···················· |
| P | 2553 | hCoV-19/Denmark/DCGC-8947/2020\|EPI_ISL_626172\|2020 | ···················· | ···················· | ···················· | ···················· | ···················· | ···················· |
| P | 2554 | hCoV-19/Denmark/DCGC-8957/2020\|EPI_ISL_626173\|2020 | ···················· | ···················· | ···················· | ···················· | ···················· | ···················· |
| P | 2555 | hCoV-19/Denmark/DCGC-8978/2020\|EPI_ISL_626174\|2020 | ···················· | ···················· | ···················· | ···················· | ···················· | ···················· |
| P | 2556 | hCoV-19/Denmark/DCGC-8986/2020\|EPI_ISL_626175\|2020 | ···················· | ···················· | ···················· | ···················· | ···················· | ···················· |
| P | 2557 | hCoV-19/Denmark/DCGC-8991/2020\|EPI_ISL_626176\|2020 | ···················· | ···················· | ···················· | ···················· | ···················· | ···················· |
| P | 2558 | hCoV-19/Denmark/DCGC-9007/2020\|EPI_ISL_626177\|2020 | ···················· | ···················· | ···················· | ···················· | ···················· | ···················· |
| P | 2559 | hCoV-19/Denmark/DCGC-8958/2020\|EPI_ISL_626178\|2020 | ···················· | ···················· | ···················· | ···················· | ···················· | ···················· |
| P | 2560 | hCoV-19/Denmark/DCGC-9000/2020\|EPI_ISL_626179\|2020 | ···················· | ···················· | ···················· | ···················· | ···················· | ···················· |
| P | 2561 | hCoV-19/Denmark/DCGC-6305/2020\|EPI_ISL_615451\|2020 | ···················· | ···················· | ···················· | ···················· | ···················· | ···················· |
| P | 2562 | hCoV-19/Denmark/DCGC-6306/2020\|EPI_ISL_615452\|2020 | ·······T············ | ···················· | ···················· | ···················· | ···················· | ···················· |
| P | 2563 | hCoV-19/Denmark/DCGC-3678/2020\|EPI_ISL_615453\|2020 | ···················· | ···················· | ···················· | ···················· | ···················· | ···················· |
| P | 2564 | hCoV-19/Denmark/DCGC-3685/2020\|EPI_ISL_615460\|2020 | ···················· | ···················· | ···················· | ···················· | ···················· | ···················· |
| P | 2565 | hCoV-19/Denmark/DCGC-3686/2020\|EPI_ISL_615461\|2020 | ···················· | ···················· | ···················· | ···················· | ···················· | ···················· |
| P | 2566 | hCoV-19/Denmark/DCGC-3688/2020\|EPI_ISL_615463\|2020 | ···················· | ···················· | ···················· | ···················· | ···················· | ···················· |
| P | 2567 | hCoV-19/Denmark/DCGC-3689/2020\|EPI_ISL_615464\|2020 | ···················· | ···················· | ···················· | ···················· | ···················· | ···················· |
| P | 2568 | hCoV-19/Denmark/DCGC-3690/2020\|EPI_ISL_615465\|2020 | ···················· | ···················· | ···················· | ···················· | ···················· | ···················· |
| P | 2569 | hCoV-19/Denmark/DCGC-3691/2020\|EPI_ISL_615466\|2020 | ···················· | ···················· | ···················· | ···················· | ···················· | ···················· |
| P | 2570 | hCoV-19/Denmark/DCGC-1724/2020\|EPI_ISL_615467\|2020 | ···················· | ···················· | ···················· | ···················· | ···················· | ···················· |
| P | 2571 | hCoV-19/Denmark/DCGC-3692/2020\|EPI_ISL_615468\|2020 | ···················· | ···················· | ···················· | ···················· | ···················· | ···················· |
| P | 2572 | hCoV-19/Denmark/DCGC-6307/2020\|EPI_ISL_615470\|2020 | ···················· | ···················· | ···················· | ···················· | ···················· | ···················· |
| P | 2573 | hCoV-19/Denmark/DCGC-3694/2020\|EPI_ISL_615471\|2020 | ···················· | ···················· | ···················· | ···················· | ···················· | ···················· |
| P | 2574 | hCoV-19/Denmark/DCGC-3695/2020\|EPI_ISL_615472\|2020 | ···················· | ···················· | ···················· | ···················· | ···················· | ···················· |
| P | 2575 | hCoV-19/Denmark/DCGC-6308/2020\|EPI_ISL_615473\|2020 | ···················· | ···················· | ···················· | ···················· | ···················· | ···················· |
| P | 2576 | hCoV-19/Denmark/DCGC-3696/2020\|EPI_ISL_615474\|2020 | ···················· | ·A·················· | ···················· | ···················· | ···················· | ··A················· |
| P | 2577 | hCoV-19/Denmark/DCGC-3697/2020\|EPI_ISL_615475\|2020 | ···················· | ···················· | ···················· | ···················· | ···················· | ···················· |
| P | 2578 | hCoV-19/Denmark/DCGC-3699/2020\|EPI_ISL_615477\|2020 | ···················· | ···················· | ···················· | ···················· | ···················· | ···················· |
| P | 2579 | hCoV-19/Denmark/DCGC-3700/2020\|EPI_ISL_615478\|2020 | ···················· | ···················· | ···················· | ···················· | ···················· | ···················· |
| P | 2580 | hCoV-19/Denmark/DCGC-3701/2020\|EPI_ISL_615479\|2020 | ···················· | ···················· | ···················· | ···················· | ···················· | ···················· |
| P | 2581 | hCoV-19/Denmark/DCGC-3702/2020\|EPI_ISL_615480\|2020 | ···················· | ···················· | ···················· | ···················· | ···················· | ···················· |
| P | 2582 | hCoV-19/Denmark/DCGC-3703/2020\|EPI_ISL_615481\|2020 | ···················· | ···················· | ···················· | ···················· | ···················· | ···················· |
| P | 2583 | hCoV-19/Denmark/DCGC-3704/2020\|EPI_ISL_615482\|2020 | ···················· | ···················· | ···················· | ···················· | ···················· | ···················· |
| P | 2584 | hCoV-19/Denmark/DCGC-3705/2020\|EPI_ISL_615483\|2020 | ···················· | ···················· | ···················· | ···················· | ···················· | ···················· |
| P | 2585 | hCoV-19/Denmark/DCGC-3706/2020\|EPI_ISL_615484\|2020 | ···················· | ·A·················· | ···················· | ···················· | ···················· | ··A················· |
| P | 2586 | hCoV-19/Denmark/DCGC-3707/2020\|EPI_ISL_615485\|2020 | ···················· | ···················· | ···················· | ···················· | ···················· | ···················· |
| P | 2587 | hCoV-19/Denmark/DCGC-3708/2020\|EPI_ISL_615486\|2020 | ···················· | ···················· | ···················· | ···················· | ···················· | ···················· |
| P | 2588 | hCoV-19/Denmark/DCGC-3709/2020\|EPI_ISL_615487\|2020 | ···················· | ···················· | ···················· | ···················· | ···················· | ···················· |
| P | 2589 | hCoV-19/Denmark/DCGC-3710/2020\|EPI_ISL_615488\|2020 | ···················· | ···················· | ···················· | ···················· | ···················· | ···················· |
| P | 2590 | hCoV-19/Denmark/DCGC-3711/2020\|EPI_ISL_615489\|2020 | ···················· | ···················· | ···················· | ···················· | ···················· | ···················· |
| P | 2591 | hCoV-19/Denmark/DCGC-3007/2020\|EPI_ISL_616783\|2020 | ·······T············ | ···················· | ···················· | ···················· | ···················· | ···················· |
| P | 2592 | hCoV-19/Denmark/DCGC-3012/2020\|EPI_ISL_616790\|2020 | ···················· | ···················· | ···················· | ···················· | ···················· | ···················· |
| P | 2593 | hCoV-19/Denmark/DCGC-3013/2020\|EPI_ISL_616791\|2020 | ···················· | ·A·················· | ···················· | ···················· | ···················· | ··A················· |
| P | 2594 | hCoV-19/Denmark/DCGC-3014/2020\|EPI_ISL_616792\|2020 | ···················· | ···················· | ···················· | ···················· | ···················· | ···················· |
| P | 2595 | hCoV-19/Denmark/DCGC-3015/2020\|EPI_ISL_616793\|2020 | ···················· | ···················· | ···················· | ···················· | ···················· | ···················· |
| P | 2596 | hCoV-19/Denmark/DCGC-3016/2020\|EPI_ISL_616794\|2020 | ···················· | ···················· | ···················· | ···················· | ···················· | ···················· |

P 2597 hCoV-19/Denmark/DCGC-3017/2020|EPI_ISL_616795|2020 ···················· ···················· ····················· ···················· ···················· ····················

P 2598 hCoV-19/Denmark/DCGC-3018/2020|EPI_ISL_616796|2020 ···················· ···················· ····················· ···················· ···················· ····················

P 2599 hCoV-19/Denmark/DCGC-3019/2020|EPI_ISL_616797|2020 ···················· ···················· ····················· ···················· ···················· ····················

P 2600 hCoV-19/Denmark/DCGC-3020/2020|EPI_ISL_616798|2020 ···················· ···················· ····················· ···················· ···················· ····················

P 2601 hCoV-19/Denmark/DCGC-3021/2020|EPI_ISL_616799|2020 ···················· ···················· ····················· ···················· ···················· ····················

P 2602 hCoV-19/Denmark/DCGC-5695/2020|EPI_ISL_621200|2020 ···················· ···················· ····················· ···················· ···················· ····················

P 2603 hCoV-19/Denmark/DCGC-5697/2020|EPI_ISL_621202|2020 ···················· ···················· ····················· ···················· ···················· ····················

P 2604 hCoV-19/Denmark/DCGC-5698/2020|EPI_ISL_621203|2020 ···················· ···················· ····················· ···················· ···················· ····················

P 2605 hCoV-19/Denmark/DCGC-5699/2020|EPI_ISL_621204|2020 ···················· ···················· ····················· ···················· ···················· ····················

P 2606 hCoV-19/Denmark/DCGC-5744/2020|EPI_ISL_621249|2020 ···················· ···················· ····················· ···················· ···················· ····················

P 2607 hCoV-19/Denmark/DCGC-5745/2020|EPI_ISL_621250|2020 ···················· ···················· ····················· ···················· ···················· ····················

P 2608 hCoV-19/Denmark/DCGC-5746/2020|EPI_ISL_621251|2020 ···················· ···················· ····················· ···················· ···················· ····················

P 2609 hCoV-19/Denmark/DCGC-5747/2020|EPI_ISL_621252|2020 ···················· ···················· ····················· ···················· ···················· ····················

P 2610 hCoV-19/Denmark/DCGC-5748/2020|EPI_ISL_621253|2020 ···················· ···················· ····················· ···················· ···················· ····················

P 2611 hCoV-19/Denmark/DCGC-5749/2020|EPI_ISL_621254|2020 ···················· ···················· ····················· ···················· ···················· ····················

P 2612 hCoV-19/Denmark/DCGC-5750/2020|EPI_ISL_621255|2020 ···················· ···················· ····················· ···················· ···················· ····················

P 2613 hCoV-19/Denmark/DCGC-5751/2020|EPI_ISL_621256|2020 ···················· ···················· ····················· ···················· ···················· ····················

P 2614 hCoV-19/Denmark/DCGC-5752/2020|EPI_ISL_621257|2020 ···················· ···················· ····················· ···················· ···················· ····················

P 2615 hCoV-19/Denmark/DCGC-5753/2020|EPI_ISL_621258|2020 ···················· ···················· ····················· ···················· ···················· ····················

P 2616 hCoV-19/Denmark/DCGC-5754/2020|EPI_ISL_621259|2020 ···················· ···················· ····················· ···················· ···················· ····················

P 2617 hCoV-19/Denmark/DCGC-5755/2020|EPI_ISL_621260|2020 ···················· ···················· ····················· ···················· ···················· ····················

P 2618 hCoV-19/Denmark/DCGC-5756/2020|EPI_ISL_621261|2020 ···················· ···················· ····················· ···················· ···················· ····················

P 2619 hCoV-19/Denmark/DCGC-4593/2020|EPI_ISL_621262|2020 ···················· ···················· ····················· ···················· ···················· ····················

P 2620 hCoV-19/Denmark/DCGC-4594/2020|EPI_ISL_621263|2020 ···················· ···················· ····················· ···················· ···················· ····················

P 2621 hCoV-19/Denmark/DCGC-4595/2020|EPI_ISL_621264|2020 ···················· ···················· ····················· ···················· ···················· ····················

P 2622 hCoV-19/Denmark/DCGC-4596/2020|EPI_ISL_621265|2020 ···················· ···················· ····················· ···················· ···················· ····················

P 2623 hCoV-19/Denmark/DCGC-4597/2020|EPI_ISL_621266|2020 ···················· ···················· ····················· ···················· ···················· ····················

P 2624 hCoV-19/Denmark/DCGC-4598/2020|EPI_ISL_621267|2020 ···················· ···················· ····················· ···················· ···················· ····················

P 2625 hCoV-19/Denmark/DCGC-4599/2020|EPI_ISL_621268|2020 ···················· ···················· ····················· ···················· ···················· ····················

P 2626 hCoV-19/Denmark/DCGC-4600/2020|EPI_ISL_621269|2020 ···················· ···················· ····················· ···················· ···················· ····················

P 2627 hCoV-19/Denmark/DCGC-4611/2020|EPI_ISL_621280|2020 ···················· ···················· ····················· ···················· ···················· ····················

P 2628 hCoV-19/Denmark/DCGC-4612/2020|EPI_ISL_621281|2020 ···················· ···················· ····················· ···················· ···················· ····················

P 2629 hCoV-19/Denmark/DCGC-4613/2020|EPI_ISL_621282|2020 ···················· ···················· ····················· ···················· ···················· ····················

P 2630 hCoV-19/Denmark/DCGC-4614/2020|EPI_ISL_621283|2020 ···················· ···················· ····················· ···················· ···················· ····················

P 2631 hCoV-19/Denmark/DCGC-4615/2020|EPI_ISL_621284|2020 ···················· ···················· ····················· ···················· ···················· ····················

P 2632 hCoV-19/Denmark/DCGC-1649/2020|EPI_ISL_622500|2020 ···················· ···················· ····················· ···················· ···················· ····················

P 2633 hCoV-19/Denmark/DCGC-1910/2020|EPI_ISL_622501|2020 ···················· ···················· ····················· ···················· ···················· ····················

P 2634 hCoV-19/Denmark/DCGC-1911/2020|EPI_ISL_622502|2020 ···················· ···················· ····················· ···················· ···T················ ····················

P 2635 hCoV-19/Denmark/DCGC-1912/2020|EPI_ISL_622503|2020 ···················· ·A·················· ····················· ···················· ···················· ····················

P 2636 hCoV-19/Denmark/DCGC-1913/2020|EPI_ISL_622504|2020 ···················· ···················· ····················· ···················· ···················· ····················

P 2637 hCoV-19/Denmark/DCGC-1914/2020|EPI_ISL_622505|2020 ···················· ···················· ····················· ···················· ···················· ····················

P 2638 hCoV-19/Denmark/DCGC-1941/2020|EPI_ISL_622506|2020 ···················· ···················· ····················· ···················· ···················· ····················

P 2639 hCoV-19/Denmark/DCGC-1915/2020|EPI_ISL_622507|2020 ···················· ···················· ····················· ···················· ···················· ····················

P 2640 hCoV-19/Denmark/DCGC-1946/2020|EPI_ISL_622508|2020 ···················· ···················· ····················· ···················· ···················· ····················

P 2641 hCoV-19/Denmark/DCGC-1916/2020|EPI_ISL_622509|2020 ···················· ···················· ····················· ···················· ···················· ····················

P 2642 hCoV-19/Denmark/DCGC-1917/2020|EPI_ISL_622510|2020 ···················· ···················· ····················· ···················· ···················· ····················

P 2643 hCoV-19/Denmark/DCGC-1918/2020|EPI_ISL_622511|2020 ···················· ···················· ····················· ···················· ···················· ····················

P 2644 hCoV-19/Denmark/DCGC-1919/2020|EPI_ISL_622512|2020 ···················· ···················· ····················· ···················· ···················· ····················

P 2645 hCoV-19/Denmark/DCGC-1920/2020|EPI_ISL_622513|2020 ···················· ···················· ····················· ···················· ···················· ····················

P 2646 hCoV-19/Denmark/DCGC-1921/2020|EPI_ISL_622514|2020 ···················· ···················· ····················· ···················· ···················· ····················

P 2647 hCoV-19/Denmark/DCGC-1923/2020|EPI_ISL_622516|2020 ···················· ···················· ····················· ···················· ···················· ····················

P 2648 hCoV-19/Denmark/DCGC-1924/2020|EPI_ISL_622517|2020 ···················· ···················· ····················· ···················· ···················· ····················

P 2649 hCoV-19/Denmark/DCGC-1925/2020|EPI_ISL_622518|2020 ···················· ···················· ····················· ···················· ···················· ····················

P 2650 hCoV-19/Denmark/DCGC-1926/2020|EPI_ISL_622519|2020 ···················· ···················· ····················· ···················· ···················· ····················

P 2651 hCoV-19/Denmark/DCGC-1927/2020|EPI_ISL_622520|2020 ···················· ·A·················· ····················· ···················· ···················· ··A·················

P 2652 hCoV-19/Denmark/DCGC-1928/2020|EPI_ISL_622521|2020 ···················· ···················· ····················· ···················· ···················· ····················

P 2653 hCoV-19/Denmark/DCGC-1929/2020|EPI_ISL_622522|2020 ···················· ···················· ····················· ···················· ···················· ····················

P 2654 hCoV-19/Denmark/DCGC-1930/2020|EPI_ISL_622523|2020 ···················· ···················· ····················· ···················· ···················· ····················

P 2655 hCoV-19/Denmark/DCGC-1650/2020|EPI_ISL_622524|2020 ···················· ···················· ····················· ···················· ···················· ····················

P 2656 hCoV-19/Denmark/DCGC-1931/2020|EPI_ISL_622525|2020 ···················· ···················· ····················· ···················· ···················· ····················

P 2657 hCoV-19/Denmark/DCGC-1932/2020|EPI_ISL_622526|2020 ···················· ···················· ····················· ···················· ···················· ····················

P 2658 hCoV-19/Denmark/DCGC-1933/2020|EPI_ISL_622527|2020 ···················· ···················· ····················· ···················· ···················· ····················

P 2659 hCoV-19/Denmark/DCGC-1934/2020|EPI_ISL_622528|2020 ···················· ···················· ····················· ···················· ···················· ····················

P 2660 hCoV-19/Denmark/DCGC-1947/2020|EPI_ISL_622529|2020 ···················· ···················· ····················· ···················· ···················· ····················

P 2661 hCoV-19/Denmark/DCGC-1935/2020|EPI_ISL_622530|2020 ···················· ···················· ····················· ···················· ···················· ····················

P 2662 hCoV-19/Denmark/DCGC-1948/2020|EPI_ISL_622531|2020 ···················· ···················· ····················· ···················· ···················· ····················

P 2663 hCoV-19/Denmark/DCGC-1936/2020|EPI_ISL_622532|2020 ···················· ···················· ····················· ···················· ···················· ····················

P 2664 hCoV-19/Denmark/DCGC-1825/2020|EPI_ISL_622533|2020 ···················· ···················· ····················· ···················· ···················· ····················

P 2665 hCoV-19/Denmark/DCGC-1833/2020|EPI_ISL_622534|2020 ········T··········· ···················· ···········T········· ···················· ···················· ····················

P 2666 hCoV-19/Denmark/DCGC-1841/2020|EPI_ISL_622535|2020 ···················· ···················· ····················· ···················· ···················· ····················

P 2667 hCoV-19/Denmark/DCGC-1871/2020|EPI_ISL_622580|2020 ···················· ···················· ····················· ···················· ···················· ····················

P 2668 hCoV-19/Denmark/DCGC-1883/2020|EPI_ISL_622581|2020 ···················· ···················· ····················· ···················· ···················· ····················

P 2669 hCoV-19/Denmark/DCGC-1897/2020|EPI_ISL_622583|2020 ···················· ···················· ····················· ···················· ···················· ····················

P 2670 hCoV-19/Denmark/DCGC-1902/2020|EPI_ISL_622584|2020 ···················· ···················· ····················· ···················· ···················· ····················

P 2671 hCoV-19/Denmark/DCGC-1830/2020|EPI_ISL_622585|2020 ···················· ···················· ····················· ···················· ···················· ····················

P 2672 hCoV-19/Denmark/DCGC-1838/2020|EPI_ISL_622586|2020 ···················· ···················· ····················· ···················· ···················· ····················

P 2673 hCoV-19/Denmark/DCGC-1845/2020|EPI_ISL_622587|2020 ···················· ···················· ····················· ···················· ···················· ····················

P 2674 hCoV-19/Denmark/DCGC-1938/2020|EPI_ISL_622588|2020 ···················· ···················· ····················· ···················· ···················· ····················

P 2675 hCoV-19/Denmark/DCGC-1858/2020|EPI_ISL_622589|2020 ···················· ···················· ····················· ···················· ···················· ····················

P 2676 hCoV-19/Denmark/DCGC-1872/2020|EPI_ISL_622590|2020 ···················· ···················· ····················· ···················· ···················· ····················

| | | | | | | | |
|---|---|---|---|---|---|---|---|
| P | 2677 hCoV-19/Denmark/DCGC-1878/2020\|EPI_ISL_622591\|2020 | ···················· | ···················· | ···················· | ···················· | ···················· | ···················· |
| P | 2678 hCoV-19/Denmark/DCGC-1891/2020\|EPI_ISL_622592\|2020 | ···················· | ···················· | ···················· | ···················· | ···················· | ···················· |
| P | 2679 hCoV-19/Denmark/DCGC-1898/2020\|EPI_ISL_622593\|2020 | ···················· | ···················· | ···················· | ···················· | ···················· | ···················· |
| P | 2680 hCoV-19/Denmark/DCGC-1831/2020\|EPI_ISL_622594\|2020 | ···················· | ···················· | ···················· | ···················· | ···················· | ···················· |
| P | 2681 hCoV-19/Denmark/DCGC-1839/2020\|EPI_ISL_622595\|2020 | ···················· | ···················· | ···················· | ···················· | ···················· | ···················· |
| P | 2682 hCoV-19/Denmark/DCGC-1846/2020\|EPI_ISL_622596\|2020 | ···················· | ···················· | ···················· | ···················· | ···················· | ···················· |
| P | 2683 hCoV-19/Denmark/DCGC-1859/2020\|EPI_ISL_622597\|2020 | ···················· | ···················· | ···················· | ···················· | ···················· | ···················· |
| P | 2684 hCoV-19/Denmark/DCGC-1873/2020\|EPI_ISL_622599\|2020 | ···················· | ···················· | ···················· | ···················· | ···················· | ···················· |
| P | 2685 hCoV-19/England/CAMC-AB14DB/2020\|EPI_ISL_623800\|20 | ···················· | ···················· | ···················· | ···················· | ···················· | ···················· |
| P | 2686 hCoV-19/England/CAMC-AB1189/2020\|EPI_ISL_623802\|20 | ···················· | ···················· | ···················· | ···················· | ···················· | ···················· |
| P | 2687 hCoV-19/England/MILK-AB8E12/2020\|EPI_ISL_623803\|20 | ···················· | ···················· | ···················· | ···················· | ···················· | ···················· |
| P | 2688 hCoV-19/England/MILK-AB7D43/2020\|EPI_ISL_623804\|20 | ···················· | ···················· | ···················· | ···················· | ···················· | ···················· |
| P | 2689 hCoV-19/England/MILK-AB7C28/2020\|EPI_ISL_623805\|20 | ···················· | ···················· | ···················· | ···················· | ···················· | ···················· |
| P | 2690 hCoV-19/England/CAMC-AB1295/2020\|EPI_ISL_623806\|20 | ···················· | ···················· | ···················· | ···················· | ···················· | ···················· |
| P | 2691 hCoV-19/England/MILK-AB754B/2020\|EPI_ISL_623807\|20 | ···················· | ···················· | ···················· | ···················· | ···················· | ···················· |
| P | 2692 hCoV-19/England/CAMC-AB130B/2020\|EPI_ISL_623808\|20 | ···················· | ···················· | ···················· | ···················· | ···················· | ···················· |
| P | 2693 hCoV-19/England/CAMC-AB18B1/2020\|EPI_ISL_623811\|20 | ···················· | ···················· | ···················· | ···················· | ···················· | ···················· |
| P | 2694 hCoV-19/England/CAMC-AB1981/2020\|EPI_ISL_623812\|20 | ···················· | ···················· | ···················· | ···················· | ···················· | ···················· |
| P | 2695 hCoV-19/England/MILK-AB88BA/2020\|EPI_ISL_623813\|20 | ···················· | ···················· | ···················· | ···················· | ···················· | ···················· |
| P | 2696 hCoV-19/England/MILK-AB795E/2020\|EPI_ISL_623814\|20 | ···················· | ···················· | ···················· | ···················· | ···················· | ···················· |
| P | 2697 hCoV-19/England/CAMC-AB2197/2020\|EPI_ISL_623815\|20 | ···················· | ···················· | ···················· | ···················· | ···················· | ···················· |
| P | 2698 hCoV-19/England/CAMC-AB1884/2020\|EPI_ISL_623816\|20 | ···················· | ···················· | ···················· | ···················· | ···················· | ···················· |
| P | 2699 hCoV-19/England/MILK-AB7D07/2020\|EPI_ISL_623817\|20 | ···················· | ···················· | ···················· | ···················· | ···················· | ···················· |
| P | 2700 hCoV-19/England/MILK-AB7D52/2020\|EPI_ISL_623818\|20 | ···················· | ···················· | ···················· | ···················· | ···················· | ···················· |
| P | 2701 hCoV-19/England/MILK-AB85C2/2020\|EPI_ISL_623819\|20 | ···················· | ·A·················· | ···················· | ···················· | ···················· | ··A················· |
| P | 2702 hCoV-19/England/MILK-AB939A/2020\|EPI_ISL_623820\|20 | ···················· | ···················· | ···················· | ···················· | ···················· | ···················· |
| P | 2703 hCoV-19/England/MILK-AB7E9B/2020\|EPI_ISL_623821\|20 | ···················· | ···················· | ···················· | ···················· | ···················· | ···················· |
| P | 2704 hCoV-19/England/CAMC-AB1383/2020\|EPI_ISL_623823\|20 | ···················· | ···················· | ···················· | ···················· | ···················· | ···················· |
| P | 2705 hCoV-19/England/MILK-AB9BB0/2020\|EPI_ISL_623824\|20 | ···················· | ···················· | ···················· | ···················· | ···················· | ···················· |
| P | 2706 hCoV-19/England/MILK-AB8D51/2020\|EPI_ISL_623825\|20 | ···················· | ···················· | ···················· | ···················· | ···················· | ···················· |
| P | 2707 hCoV-19/England/MILK-AB7CBF/2020\|EPI_ISL_623826\|20 | ···················· | ···················· | ···················· | ···················· | ···················· | ···················· |
| P | 2708 hCoV-19/England/CAMC-AB1B9A/2020\|EPI_ISL_623829\|20 | ···················· | ···················· | ···················· | ···················· | ···················· | ···················· |
| P | 2709 hCoV-19/England/CAMC-AB116B/2020\|EPI_ISL_623831\|20 | ···················· | ···················· | ···················· | ···················· | ···················· | ···················· |
| P | 2710 hCoV-19/England/CAMC-AB15AB/2020\|EPI_ISL_623832\|20 | ···················· | ···················· | ···················· | ···················· | ···················· | ···················· |
| P | 2711 hCoV-19/England/CAMC-AB1848/2020\|EPI_ISL_623833\|20 | ···················· | ···················· | ···················· | ···················· | ···················· | ···················· |
| P | 2712 hCoV-19/England/CAMC-AB1AF7/2020\|EPI_ISL_623834\|20 | ···················· | ···················· | ···················· | ···················· | ···················· | ···················· |
| P | 2713 hCoV-19/England/MILK-AB936D/2020\|EPI_ISL_623835\|20 | ···················· | ···················· | ···················· | ···················· | ···················· | ···················· |
| P | 2714 hCoV-19/England/MILK-AB79C7/2020\|EPI_ISL_623836\|20 | ···················· | ···················· | ···················· | ···················· | ···················· | ···················· |
| P | 2715 hCoV-19/England/MILK-AB7A00/2020\|EPI_ISL_623837\|20 | ···················· | ···················· | ···················· | ···················· | ···················· | ···················· |
| P | 2716 hCoV-19/England/CAMC-AB1374/2020\|EPI_ISL_623838\|20 | ···················· | ···················· | ···················· | ···················· | ···················· | ···················· |
| P | 2717 hCoV-19/England/CAMC-AB2610/2020\|EPI_ISL_623841\|20 | ···················· | ···················· | ···················· | ···················· | ···················· | ···················· |
| P | 2718 hCoV-19/England/MILK-AB755A/2020\|EPI_ISL_623842\|20 | ···················· | ···················· | ···················· | ···················· | ···················· | ···················· |
| P | 2719 hCoV-19/England/CAMC-AB1F07/2020\|EPI_ISL_623844\|20 | ···················· | ···················· | ···················· | ···················· | ···················· | ···················· |
| P | 2720 hCoV-19/England/MILK-AB88D8/2020\|EPI_ISL_623845\|20 | ···················· | ···················· | ···················· | ···················· | ·····T·············· | ···················· |
| P | 2721 hCoV-19/England/CAMC-AB1EFB/2020\|EPI_ISL_623846\|20 | ···················· | ···················· | ···················· | ···················· | ···················· | ···················· |
| P | 2722 hCoV-19/England/CAMC-AB1ECE/2020\|EPI_ISL_623847\|20 | ···················· | ···················· | ···················· | ···················· | ···················· | ···················· |
| P | 2723 hCoV-19/England/CAMC-AB1972/2020\|EPI_ISL_623849\|20 | ···················· | ···················· | ···················· | ···················· | ···················· | ···················· |
| P | 2724 hCoV-19/England/MILK-AB938B/2020\|EPI_ISL_623851\|20 | ···················· | ···················· | ···················· | ···················· | ···················· | ···················· |
| P | 2725 hCoV-19/England/MILK-AB8D60/2020\|EPI_ISL_623852\|20 | ···················· | ···················· | ···················· | ···················· | ···················· | ···················· |
| P | 2726 hCoV-19/England/MILK-AB74F3/2020\|EPI_ISL_623853\|20 | ···················· | ·A·················· | ···················· | ···················· | ···················· | ··A················· |
| P | 2727 hCoV-19/England/CAMC-AB1E91/2020\|EPI_ISL_623854\|20 | ···················· | ·A·················· | ···················· | ···················· | ···················· | ···················· |
| P | 2728 hCoV-19/England/CAMC-AB1D58/2020\|EPI_ISL_623856\|20 | ···················· | ···················· | ···················· | ···················· | ···················· | ···················· |
| P | 2729 hCoV-19/England/CAMC-AB21C4/2020\|EPI_ISL_623857\|20 | ···················· | ···················· | ···················· | ···················· | ···················· | ···················· |
| P | 2730 hCoV-19/England/ALDP-AA0E68/2020\|EPI_ISL_623858\|20 | ···················· | ···················· | ···················· | ···················· | ···················· | ···················· |
| P | 2731 hCoV-19/England/ALDP-AA2F45/2020\|EPI_ISL_623859\|20 | ···················· | ···················· | ···················· | ···················· | ···················· | ···················· |
| P | 2732 hCoV-19/England/ALDP-AA141A/2020\|EPI_ISL_623860\|20 | ···················· | ···················· | ···················· | ···················· | ···················· | ···················· |
| P | 2733 hCoV-19/England/ALDP-AA19C0/2020\|EPI_ISL_623862\|20 | ···················· | ···················· | ···················· | ···················· | ···················· | ···················· |
| P | 2734 hCoV-19/Scotland/QEUH-AAD320/2020\|EPI_ISL_623863\|2 | ···················· | ···················· | ···················· | ···················· | ···················· | ···················· |
| P | 2735 hCoV-19/England/QEUH-AAEF49/2020\|EPI_ISL_623865\|20 | ···················· | ···················· | ···················· | ···················· | ···················· | ···················· |
| P | 2736 hCoV-19/England/ALDP-AA2F54/2020\|EPI_ISL_623866\|20 | ···················· | ···················· | ···················· | ···················· | ···················· | ···················· |
| P | 2737 hCoV-19/Denmark/DCGC-8747/2020\|EPI_ISL_626060\|2020 | ···················· | ···················· | ···················· | ···················· | ···················· | ···················· |
| P | 2738 hCoV-19/Denmark/DCGC-8749/2020\|EPI_ISL_626061\|2020 | ···················· | ···················· | ···················· | ···················· | ···················· | ···················· |
| P | 2739 hCoV-19/Denmark/DCGC-8755/2020\|EPI_ISL_626062\|2020 | ···················· | ···················· | ···················· | ···················· | ···················· | ···················· |
| P | 2740 hCoV-19/Denmark/DCGC-8765/2020\|EPI_ISL_626063\|2020 | ···················· | ···················· | ···················· | ···················· | ···················· | ···················· |
| P | 2741 hCoV-19/Denmark/DCGC-8773/2020\|EPI_ISL_626064\|2020 | ···················· | ···················· | ···················· | ···················· | ···················· | ···················· |
| P | 2742 hCoV-19/Denmark/DCGC-8790/2020\|EPI_ISL_626065\|2020 | ···················· | ···················· | ···················· | ···················· | ···················· | ···················· |
| P | 2743 hCoV-19/Denmark/DCGC-8798/2020\|EPI_ISL_626066\|2020 | ···················· | ···················· | ···················· | ···················· | ···················· | ···················· |
| P | 2744 hCoV-19/Denmark/DCGC-8748/2020\|EPI_ISL_626067\|2020 | ···················· | ·A·················· | ···················· | ···················· | ···················· | ··A················· |
| P | 2745 hCoV-19/Denmark/DCGC-8756/2020\|EPI_ISL_626068\|2020 | ···················· | ···················· | ···················· | ···················· | ···················· | ···················· |
| P | 2746 hCoV-19/Denmark/DCGC-8766/2020\|EPI_ISL_626069\|2020 | ···················· | ···················· | ···················· | ···················· | ···················· | ···················· |
| P | 2747 hCoV-19/Denmark/DCGC-8799/2020\|EPI_ISL_626070\|2020 | ···················· | ···················· | ···················· | ···················· | ···················· | ···················· |
| P | 2748 hCoV-19/Denmark/DCGC-8762/2020\|EPI_ISL_626071\|2020 | ···················· | ···················· | ···················· | ···················· | ···················· | ···················· |
| P | 2749 hCoV-19/Denmark/DCGC-8767/2020\|EPI_ISL_626072\|2020 | ···················· | ···················· | ···················· | ···················· | ···················· | ···················· |
| P | 2750 hCoV-19/Denmark/DCGC-8787/2020\|EPI_ISL_626073\|2020 | ···················· | ···················· | ···················· | ···················· | ···················· | ···················· |
| P | 2751 hCoV-19/Denmark/DCGC-8792/2020\|EPI_ISL_626074\|2020 | ···················· | ···················· | ···················· | ···················· | ···················· | ···················· |
| P | 2752 hCoV-19/Denmark/DCGC-8800/2020\|EPI_ISL_626075\|2020 | ···················· | ···················· | ···················· | ···················· | ···················· | ···················· |
| P | 2753 hCoV-19/Denmark/DCGC-8812/2020\|EPI_ISL_626076\|2020 | ···················· | ···················· | ···················· | ···················· | ···················· | ···················· |
| P | 2754 hCoV-19/Denmark/DCGC-8819/2020\|EPI_ISL_626077\|2020 | ···················· | ···················· | ···················· | ···················· | ···················· | ···················· |
| P | 2755 hCoV-19/Denmark/DCGC-8826/2020\|EPI_ISL_626078\|2020 | ···················· | ···················· | ···················· | ···················· | ···················· | ···················· |
| P | 2756 hCoV-19/Denmark/DCGC-8836/2020\|EPI_ISL_626079\|2020 | ···················· | ···················· | ···················· | ···················· | ···················· | ···················· |

```
P    2757 hCoV-19/Denmark/DCGC-8846/2020|EPI_ISL_626080|2020    ····················  ·A··················  ·····················  ····················  ····················  ····················
P    2758 hCoV-19/Denmark/DCGC-8852/2020|EPI_ISL_626081|2020    ····················  ····A···············  ·····················  ····················  ····················  ····················
P    2759 hCoV-19/Denmark/DCGC-8858/2020|EPI_ISL_626082|2020    ····················  ····················  ·····················  ····················  ····················  ····················
P    2760 hCoV-19/Denmark/DCGC-8813/2020|EPI_ISL_626083|2020    ····················  ····················  ·····················  ····················  ····················  ····················
P    2761 hCoV-19/Denmark/DCGC-8820/2020|EPI_ISL_626084|2020    ····················  ····················  ·····················  ····················  ····················  ····················
P    2762 hCoV-19/Denmark/DCGC-8827/2020|EPI_ISL_626085|2020    ····················  ····················  ·····················  ····················  ····················  ····················
P    2763 hCoV-19/Denmark/DCGC-8847/2020|EPI_ISL_626086|2020    ····················  ····················  ·····················  ····················  ····················  ····················
P    2764 hCoV-19/Denmark/DCGC-8853/2020|EPI_ISL_626087|2020    ····················  ····A···············  ·····················  ····················  ····················  ····················
P    2765 hCoV-19/Denmark/DCGC-8859/2020|EPI_ISL_626088|2020    ····················  ····················  ·····················  ····················  ····················  ····················
P    2766 hCoV-19/Denmark/DCGC-8807/2020|EPI_ISL_626089|2020    ····················  ····················  ·····················  ····················  ····················  ····················
P    2767 hCoV-19/Denmark/DCGC-8814/2020|EPI_ISL_626090|2020    ····················  ····················  ·····················  ····················  ····················  ····················
P    2768 hCoV-19/Denmark/DCGC-8821/2020|EPI_ISL_626091|2020    ····················  ····················  ·····················  ····················  ····················  ····················
P    2769 hCoV-19/Denmark/DCGC-8831/2020|EPI_ISL_626092|2020    ····················  ····················  ·····················  ····················  ····················  ····················
P    2770 hCoV-19/Denmark/DCGC-8848/2020|EPI_ISL_626093|2020    ····················  ····················  ·····················  ····················  ····················  ····················
P    2771 hCoV-19/Denmark/DCGC-8871/2020|EPI_ISL_626094|2020    ····················  ····················  ·····················  ····················  ····················  ····················
P    2772 hCoV-19/Denmark/DCGC-8815/2020|EPI_ISL_626095|2020    ····················  ····················  ·····················  ····················  ····················  ····················
P    2773 hCoV-19/Denmark/DCGC-8822/2020|EPI_ISL_626096|2020    ····················  ····················  ·····················  ····················  ····················  ····················
P    2774 hCoV-19/Denmark/DCGC-8832/2020|EPI_ISL_626097|2020    ····················  ····················  ·····················  ····················  ····················  ····················
P    2775 hCoV-19/Denmark/DCGC-8841/2020|EPI_ISL_626098|2020    ····················  ····················  ·····················  ····················  ····················  ····················
P    2776 hCoV-19/Denmark/DCGC-8849/2020|EPI_ISL_626099|2020    ····················  ····················  ·····················  ····················  ····················  ····················
P    2777 hCoV-19/Denmark/DCGC-8854/2020|EPI_ISL_626100|2020    ····················  ····················  ·····················  ····················  ····················  ····················
P    2778 hCoV-19/Denmark/DCGC-8867/2020|EPI_ISL_626101|2020    ····················  ····················  ·····················  ····················  ····················  ····················
P    2779 hCoV-19/Denmark/DCGC-8809/2020|EPI_ISL_626102|2020    ····················  ····················  ·····················  ····················  ····················  ····················
P    2780 hCoV-19/Denmark/DCGC-8816/2020|EPI_ISL_626103|2020    ····················  ····················  ·····················  ····················  ····················  ····················
P    2781 hCoV-19/Denmark/DCGC-8823/2020|EPI_ISL_626104|2020    ····················  ·A··················  ·····················  ····················  ····················  ···A················
P    2782 hCoV-19/Denmark/DCGC-8850/2020|EPI_ISL_626105|2020    ····················  ····················  ·····················  ····················  ····················  ····················
P    2783 hCoV-19/Denmark/DCGC-8824/2020|EPI_ISL_626106|2020    ····················  ····················  ·····················  ····················  ····················  ····················
P    2784 hCoV-19/Denmark/DCGC-8834/2020|EPI_ISL_626107|2020    ····················  ····················  ·····················  ····················  ····················  ····················
P    2785 hCoV-19/Denmark/DCGC-8862/2020|EPI_ISL_626108|2020    ····················  ····················  ·····················  ····················  ····················  ····················
P    2786 hCoV-19/Denmark/DCGC-8868/2020|EPI_ISL_626109|2020    ····················  ····················  ·····················  ····················  ····················  ····················
P    2787 hCoV-19/Denmark/DCGC-8810/2020|EPI_ISL_626110|2020    ····················  ····················  ·····················  ····················  ····················  ····················
P    2788 hCoV-19/Denmark/DCGC-8817/2020|EPI_ISL_626111|2020    ····················  ····················  ·····················  ····················  ····················  ····················
P    2789 hCoV-19/Denmark/DCGC-8825/2020|EPI_ISL_626112|2020    ····················  ·A··················  ·····················  ····················  ····················  ····················
P    2790 hCoV-19/Denmark/DCGC-8835/2020|EPI_ISL_626113|2020    ····················  ····················  ·····················  ····················  ····················  ····················
P    2791 hCoV-19/Denmark/DCGC-8844/2020|EPI_ISL_626114|2020    ····················  ····················  ·····················  ····················  ····················  ····················
P    2792 hCoV-19/Denmark/DCGC-8851/2020|EPI_ISL_626115|2020    ····················  ····················  ·····················  ····················  ····················  ····················
P    2793 hCoV-19/Denmark/DCGC-8856/2020|EPI_ISL_626116|2020    ····················  ····················  ·····················  ····················  ····················  ····················
P    2794 hCoV-19/Denmark/DCGC-8863/2020|EPI_ISL_626117|2020    ····················  ····················  ·····················  ····················  ····················  ····················
P    2795 hCoV-19/Denmark/DCGC-8818/2020|EPI_ISL_626118|2020    ····················  ····················  ·····················  ····················  ····················  ····················
P    2796 hCoV-19/Denmark/DCGC-8838/2020|EPI_ISL_626119|2020    ····················  ····················  ·····················  ····················  ····················  ····················
P    2797 hCoV-19/Denmark/DCGC-8845/2020|EPI_ISL_626120|2020    ····················  ·A··················  ·····················  ····················  ····················  ···A················
P    2798 hCoV-19/Denmark/DCGC-8857/2020|EPI_ISL_626121|2020    ····················  ····················  ·····················  ····················  ····················  ····················
P    2799 hCoV-19/Denmark/DCGC-8864/2020|EPI_ISL_626122|2020    ····················  ····A···············  ·····················  ····················  ····················  ····················
P    2800 hCoV-19/Denmark/DCGC-8880/2020|EPI_ISL_626123|2020    ····················  ····················  ·····················  ····················  ····················  ····················
P    2801 hCoV-19/Denmark/DCGC-8894/2020|EPI_ISL_626124|2020    ····················  ····················  ·····················  ····················  ····················  ····················
P    2802 hCoV-19/Denmark/DCGC-8899/2020|EPI_ISL_626125|2020    ····················  ····················  ·····················  ····················  ····················  ····················
P    2803 hCoV-19/Denmark/DCGC-8907/2020|EPI_ISL_626126|2020    ····················  ····················  ·····················  ····················  ····················  ····················
P    2804 hCoV-19/Denmark/DCGC-8915/2020|EPI_ISL_626127|2020    ····················  ····················  ·····················  ····················  ····················  ····················
P    2805 hCoV-19/Denmark/DCGC-8930/2020|EPI_ISL_626128|2020    ····················  ····················  ·····················  ····················  ····················  ····················
P    2806 hCoV-19/Denmark/DCGC-8937/2020|EPI_ISL_626129|2020    ····················  ····················  ·····················  ····················  ····················  ····················
P    2807 hCoV-19/Denmark/DCGC-8874/2020|EPI_ISL_626130|2020    ····················  ····················  ·····················  ····················  ····················  ····················
P    2808 hCoV-19/Denmark/DCGC-8889/2020|EPI_ISL_626131|2020    ····················  A···················  ·····················  ····················  ····················  ····················
P    2809 hCoV-19/Denmark/DCGC-8895/2020|EPI_ISL_626132|2020    ····················  ····················  ·····················  ····················  ······T·············  ····················
P    2810 hCoV-19/Denmark/DCGC-8916/2020|EPI_ISL_626133|2020    ····················  ····················  ·····················  ····················  ····················  ····················
P    2811 hCoV-19/Denmark/DCGC-8931/2020|EPI_ISL_626134|2020    ····················  ····················  ·····················  ····················  ····················  ····················
P    2812 hCoV-19/Denmark/DCGC-8875/2020|EPI_ISL_626135|2020    ····················  ····················  ·····················  ····················  ····················  ····················
P    2813 hCoV-19/Denmark/DCGC-8882/2020|EPI_ISL_626136|2020    ····················  ····················  ·····················  ····················  ····················  ····················
P    2814 hCoV-19/Denmark/DCGC-8890/2020|EPI_ISL_626137|2020    ····················  ····················  ·····················  ····················  ····················  ····················
P    2815 hCoV-19/Denmark/DCGC-8939/2020|EPI_ISL_626138|2020    ····················  ····················  ·····················  ····················  ····················  ····················
P    2816 hCoV-19/Denmark/DCGC-8876/2020|EPI_ISL_626139|2020    ····················  ····················  ·····················  ····················  ····················  ····················
P    2817 hCoV-19/Denmark/DCGC-5371/2020|EPI_ISL_615302|2020    ····················  ····················  ·····················  ····················  ····················  ····················
P    2818 hCoV-19/Denmark/DCGC-5372/2020|EPI_ISL_615303|2020    ····················  ····················  ·····················  ····················  ····················  ····················
P    2819 hCoV-19/Denmark/DCGC-5373/2020|EPI_ISL_615304|2020    ····················  ····················  ·····················  ····················  ····················  ····················
P    2820 hCoV-19/Denmark/DCGC-5374/2020|EPI_ISL_615305|2020    ····················  ····················  ·····················  ····················  ····················  ····················
P    2821 hCoV-19/Denmark/DCGC-5375/2020|EPI_ISL_615306|2020    ····················  ····················  ·····················  ····················  ····················  ····················
P    2822 hCoV-19/Denmark/DCGC-5376/2020|EPI_ISL_615307|2020    ····················  ····················  ·····················  ····················  ····················  ····················
P    2823 hCoV-19/Denmark/DCGC-5377/2020|EPI_ISL_615308|2020    ····················  ····················  ·····················  ····················  ····················  ····················
P    2824 hCoV-19/Denmark/DCGC-5378/2020|EPI_ISL_615309|2020    ····················  ····················  ·····················  ····················  ····················  ····················
P    2825 hCoV-19/Denmark/DCGC-5379/2020|EPI_ISL_615310|2020    ····················  ····················  ·····················  ····················  ····················  ····················
P    2826 hCoV-19/Denmark/DCGC-5380/2020|EPI_ISL_615311|2020    ····················  ····················  ·····················  ····················  ····················  ····················
P    2827 hCoV-19/Denmark/DCGC-5381/2020|EPI_ISL_615312|2020    ····················  ····················  ·····················  ····················  ····················  ····················
P    2828 hCoV-19/Denmark/DCGC-5382/2020|EPI_ISL_615313|2020    ····················  ····················  ·····················  ····················  ····················  ····················
P    2829 hCoV-19/Denmark/DCGC-5384/2020|EPI_ISL_615315|2020    ····················  ····················  ·····················  ····················  ····················  ····················
P    2830 hCoV-19/Denmark/DCGC-5226/2020|EPI_ISL_615316|2020    ····················  ····················  ·····················  ····················  ····················  ····················
P    2831 hCoV-19/Denmark/DCGC-5385/2020|EPI_ISL_615317|2020    ····················  ····················  ·····················  ····················  ····················  ····················
P    2832 hCoV-19/Denmark/DCGC-5387/2020|EPI_ISL_615319|2020    ····················  ····················  ·····················  ····················  ····················  ····················
P    2833 hCoV-19/Denmark/DCGC-5388/2020|EPI_ISL_615320|2020    ····················  ····················  ·····················  ····················  ····················  ····················
P    2834 hCoV-19/Denmark/DCGC-5390/2020|EPI_ISL_615322|2020    ····················  ·A··················  ·····················  ····················  ····················  ···A················
P    2835 hCoV-19/Denmark/DCGC-5391/2020|EPI_ISL_615323|2020    ····················  ····················  ·····················  ····················  ····················  ····················
P    2836 hCoV-19/Denmark/DCGC-5392/2020|EPI_ISL_615324|2020    ····················  ····················  ·····················  ····················  ····················  ····················
```

| | | | | | | | |
|---|---|---|---|---|---|---|---|
| P | 2837 hCoV-19/Denmark/DCGC-5393/2020\|EPI_ISL_615325\|2020 | ···················· | ···················· | ···················· | ···················· | ···················· | ···················· |
| P | 2838 hCoV-19/Denmark/DCGC-5394/2020\|EPI_ISL_615326\|2020 | ···················· | ···················· | ···················· | ···················· | ···················· | ···················· |
| P | 2839 hCoV-19/Denmark/DCGC-5395/2020\|EPI_ISL_615327\|2020 | ···················· | ···················· | ···················· | ···················· | ···················· | ···················· |
| P | 2840 hCoV-19/Denmark/DCGC-5396/2020\|EPI_ISL_615328\|2020 | ···················· | ···················· | ···················· | ···················· | ···················· | ···················· |
| P | 2841 hCoV-19/Denmark/DCGC-5397/2020\|EPI_ISL_615329\|2020 | ···················· | ···················· | ···················· | ···················· | ···················· | ···················· |
| P | 2842 hCoV-19/Denmark/DCGC-2783/2020\|EPI_ISL_616618\|2020 | ···················· | ···················· | ···················· | ···················· | ···················· | ···················· |
| P | 2843 hCoV-19/Denmark/DCGC-2784/2020\|EPI_ISL_616619\|2020 | ···················· | ···················· | ···················· | ···················· | ···················· | ···················· |
| P | 2844 hCoV-19/Denmark/DCGC-2785/2020\|EPI_ISL_616620\|2020 | ···················· | ···················· | ···················· | ···················· | ···················· | ···················· |
| P | 2845 hCoV-19/Denmark/DCGC-2786/2020\|EPI_ISL_616621\|2020 | ···················· | ···················· | ···················· | ···················· | ···················· | ···················· |
| P | 2846 hCoV-19/Denmark/DCGC-2787/2020\|EPI_ISL_616622\|2020 | ···················· | ···················· | ···················· | ···················· | ···················· | ···················· |
| P | 2847 hCoV-19/Denmark/DCGC-2788/2020\|EPI_ISL_616623\|2020 | ···················· | ···················· | ···················· | ···················· | ···················· | ···················· |
| P | 2848 hCoV-19/Denmark/DCGC-2789/2020\|EPI_ISL_616624\|2020 | ···················· | ···················· | ···················· | ···················· | ···················· | ···················· |
| P | 2849 hCoV-19/Denmark/DCGC-2790/2020\|EPI_ISL_616625\|2020 | ···················· | ···················· | ···················· | ···················· | ···················· | ···················· |
| P | 2850 hCoV-19/Denmark/DCGC-2791/2020\|EPI_ISL_616626\|2020 | ···················· | ···················· | ···················· | ···················· | ···················· | ···················· |
| P | 2851 hCoV-19/Denmark/DCGC-2792/2020\|EPI_ISL_616627\|2020 | ···················· | ···················· | ···················· | ···················· | ···················· | ···················· |
| P | 2852 hCoV-19/Denmark/DCGC-2793/2020\|EPI_ISL_616628\|2020 | ···················· | ···················· | ···················· | ···················· | ···················· | ···················· |
| P | 2853 hCoV-19/Denmark/DCGC-2794/2020\|EPI_ISL_616629\|2020 | ···················· | ·A·················· | ···················· | ···················· | ···················· | ··A················· |
| P | 2854 hCoV-19/Denmark/DCGC-2795/2020\|EPI_ISL_616630\|2020 | ···················· | ···················· | ···················· | ···················· | ···················· | ···················· |
| P | 2855 hCoV-19/Denmark/DCGC-2796/2020\|EPI_ISL_616631\|2020 | ···················· | ···················· | ···················· | ···················· | ···················· | ···················· |
| P | 2856 hCoV-19/Denmark/DCGC-2926/2020\|EPI_ISL_616632\|2020 | ···················· | ···················· | ···················· | ···················· | ···················· | ···················· |
| P | 2857 hCoV-19/Denmark/DCGC-2797/2020\|EPI_ISL_616633\|2020 | ···················· | ···················· | ···················· | ···················· | ···················· | ···················· |
| P | 2858 hCoV-19/Denmark/DCGC-2798/2020\|EPI_ISL_616634\|2020 | ···················· | ···················· | ···················· | ···················· | ···················· | ···················· |
| P | 2859 hCoV-19/Denmark/DCGC-2799/2020\|EPI_ISL_616635\|2020 | ···················· | ···················· | ···················· | ···················· | ···················· | ···················· |
| P | 2860 hCoV-19/Denmark/DCGC-2800/2020\|EPI_ISL_616636\|2020 | ···················· | ···················· | ···················· | ···················· | ···················· | ···················· |
| P | 2861 hCoV-19/Denmark/DCGC-2802/2020\|EPI_ISL_616638\|2020 | ···················· | ···················· | ···················· | ···················· | ···················· | ···················· |
| P | 2862 hCoV-19/Denmark/DCGC-2803/2020\|EPI_ISL_616639\|2020 | ···················· | ···················· | ···················· | ···················· | ···················· | ···················· |
| P | 2863 hCoV-19/Denmark/DCGC-2804/2020\|EPI_ISL_616640\|2020 | ···················· | ···················· | ···················· | ···················· | ···················· | ···················· |
| P | 2864 hCoV-19/Denmark/DCGC-2805/2020\|EPI_ISL_616641\|2020 | ···················· | ···················· | ···················· | ···················· | ···················· | ···················· |
| P | 2865 hCoV-19/Denmark/DCGC-2806/2020\|EPI_ISL_616642\|2020 | ···················· | ···················· | ···················· | ···················· | ···················· | ···················· |
| P | 2866 hCoV-19/Denmark/DCGC-2807/2020\|EPI_ISL_616643\|2020 | ···················· | ···················· | ···················· | ···················· | ···················· | ···················· |
| P | 2867 hCoV-19/Denmark/DCGC-2808/2020\|EPI_ISL_616644\|2020 | ···················· | ···················· | ···················· | ···················· | ···················· | ···················· |
| P | 2868 hCoV-19/Denmark/DCGC-2809/2020\|EPI_ISL_616645\|2020 | ···················· | ···················· | ···················· | ···················· | ···················· | ···················· |
| P | 2869 hCoV-19/Denmark/DCGC-2810/2020\|EPI_ISL_616646\|2020 | ···················· | ···················· | ···················· | ···················· | ···················· | ···················· |
| P | 2870 hCoV-19/Denmark/DCGC-2813/2020\|EPI_ISL_616649\|2020 | ···················· | ···················· | ···················· | ···················· | ···················· | ···················· |
| P | 2871 hCoV-19/Denmark/DCGC-2814/2020\|EPI_ISL_616650\|2020 | ···················· | ·A·················· | ···················· | ···················· | ···················· | ···················· |
| P | 2872 hCoV-19/Denmark/DCGC-2927/2020\|EPI_ISL_616651\|2020 | ···················· | ···················· | ···················· | ···················· | ···················· | ···················· |
| P | 2873 hCoV-19/Denmark/DCGC-2826/2020\|EPI_ISL_616652\|2020 | ···················· | ···················· | ···················· | ···················· | ···················· | ···················· |
| P | 2874 hCoV-19/Denmark/DCGC-2827/2020\|EPI_ISL_616653\|2020 | ···················· | ···················· | ···················· | ···················· | ···················· | ···················· |
| P | 2875 hCoV-19/Denmark/DCGC-2828/2020\|EPI_ISL_616654\|2020 | ···················· | ···················· | ···················· | ···················· | ···················· | ···················· |
| P | 2876 hCoV-19/Denmark/DCGC-2829/2020\|EPI_ISL_616655\|2020 | ···················· | ···················· | ···················· | ···················· | ···················· | ···················· |
| P | 2877 hCoV-19/Denmark/DCGC-2830/2020\|EPI_ISL_616656\|2020 | ···················· | ···················· | ···················· | ···················· | ···················· | ···················· |
| P | 2878 hCoV-19/Denmark/DCGC-2831/2020\|EPI_ISL_616657\|2020 | ···················· | ···················· | ···················· | ···················· | ···················· | ···················· |
| P | 2879 hCoV-19/Denmark/DCGC-2832/2020\|EPI_ISL_616658\|2020 | ···················· | ···················· | ···················· | ···················· | ···················· | ···················· |
| P | 2880 hCoV-19/Denmark/DCGC-2833/2020\|EPI_ISL_616659\|2020 | ···················· | ···················· | ···················· | ···················· | ···················· | ···················· |
| P | 2881 hCoV-19/Denmark/DCGC-2574/2020\|EPI_ISL_617905\|2020 | ···················· | ···················· | ···················· | ···················· | ···················· | ···················· |
| P | 2882 hCoV-19/Denmark/DCGC-8412/2020\|EPI_ISL_617906\|2020 | ···················· | ···················· | ···················· | ···················· | ···················· | ···················· |
| P | 2883 hCoV-19/Denmark/DCGC-8430/2020\|EPI_ISL_617908\|2020 | ···················· | ···················· | ···················· | ···················· | ···················· | ···················· |
| P | 2884 hCoV-19/Denmark/DCGC-8437/2020\|EPI_ISL_617909\|2020 | ···················· | ···················· | ···················· | ···················· | ···················· | ···················· |
| P | 2885 hCoV-19/Denmark/DCGC-8444/2020\|EPI_ISL_617910\|2020 | ···················· | ···················· | ···················· | ···················· | ···················· | ···················· |
| P | 2886 hCoV-19/Denmark/DCGC-8450/2020\|EPI_ISL_617911\|2020 | ···················· | ···················· | ···················· | ···················· | ···················· | ···················· |
| P | 2887 hCoV-19/Denmark/DCGC-8458/2020\|EPI_ISL_617912\|2020 | ···················· | ···················· | ···················· | ···················· | ···················· | ···················· |
| P | 2888 hCoV-19/Denmark/DCGC-8464/2020\|EPI_ISL_617913\|2020 | ···················· | ·A·················· | ···················· | ···················· | ···················· | ···················· |
| P | 2889 hCoV-19/Denmark/DCGC-8470/2020\|EPI_ISL_617914\|2020 | ···················· | ···················· | ···················· | ···················· | ···················· | ···················· |
| P | 2890 hCoV-19/Denmark/DCGC-8413/2020\|EPI_ISL_617915\|2020 | ···················· | ···················· | ···················· | ···················· | ···················· | ···················· |
| P | 2891 hCoV-19/Denmark/DCGC-8419/2020\|EPI_ISL_617916\|2020 | ···················· | ···················· | ···················· | ···················· | ···················· | ···················· |
| P | 2892 hCoV-19/Denmark/DCGC-8438/2020\|EPI_ISL_617917\|2020 | ···················· | ···················· | ···················· | ···················· | ···················· | ···················· |
| P | 2893 hCoV-19/Denmark/DCGC-8459/2020\|EPI_ISL_617920\|2020 | ···················· | ···················· | ···················· | ···················· | ···················· | ···················· |
| P | 2894 hCoV-19/Denmark/DCGC-8471/2020\|EPI_ISL_617921\|2020 | ···················· | ···················· | ···················· | ···················· | ···················· | ···················· |
| P | 2895 hCoV-19/Denmark/DCGC-8414/2020\|EPI_ISL_617922\|2020 | ···················· | ···················· | ···················· | ···················· | ···················· | ···················· |
| P | 2896 hCoV-19/Denmark/DCGC-8420/2020\|EPI_ISL_617923\|2020 | ···················· | ···················· | ···················· | ···················· | ···················· | ···················· |
| P | 2897 hCoV-19/Denmark/DCGC-8439/2020\|EPI_ISL_617926\|2020 | ···················· | ···················· | ···················· | ···················· | ···················· | ···················· |
| P | 2898 hCoV-19/Denmark/DCGC-1730/2020\|EPI_ISL_617949\|2020 | ···················· | ···················· | ···················· | ···················· | ···················· | ···················· |
| P | 2899 hCoV-19/Denmark/DCGC-1731/2020\|EPI_ISL_617950\|2020 | ···················· | ···················· | ···················· | ···················· | ···················· | ···················· |
| P | 2900 hCoV-19/Denmark/DCGC-1547/2020\|EPI_ISL_617951\|2020 | ···················· | ···················· | ···················· | ···················· | ···················· | ···················· |
| P | 2901 hCoV-19/Denmark/DCGC-1546/2020\|EPI_ISL_617952\|2020 | ···················· | ···················· | ···················· | ···················· | ···················· | ···················· |
| P | 2902 hCoV-19/Denmark/DCGC-1812/2020\|EPI_ISL_617954\|2020 | ···················· | ···················· | ···················· | ···················· | ···················· | ···················· |
| P | 2903 hCoV-19/Denmark/DCGC-1289/2020\|EPI_ISL_617955\|2020 | ···················· | ···················· | ···················· | ···················· | ···················· | ···················· |
| P | 2904 hCoV-19/Denmark/DCGC-1813/2020\|EPI_ISL_617956\|2020 | ···················· | ···················· | ···················· | ···················· | ···················· | ···················· |
| P | 2905 hCoV-19/Denmark/DCGC-1290/2020\|EPI_ISL_617957\|2020 | ···················· | ···················· | ···················· | ···················· | ···················· | ···················· |
| P | 2906 hCoV-19/Denmark/DCGC-1814/2020\|EPI_ISL_617958\|2020 | ···················· | ···················· | ···················· | ···················· | ···················· | ···················· |
| P | 2907 hCoV-19/Denmark/DCGC-1541/2020\|EPI_ISL_617962\|2020 | ···················· | ···················· | ···················· | ···················· | ···················· | ···················· |
| P | 2908 hCoV-19/Denmark/DCGC-1542/2020\|EPI_ISL_617963\|2020 | ···················· | ···················· | ···················· | ···················· | ···················· | ···················· |
| P | 2909 hCoV-19/Denmark/DCGC-1543/2020\|EPI_ISL_617964\|2020 | ···················· | ···················· | ···················· | ···················· | ···················· | ···················· |
| P | 2910 hCoV-19/Denmark/DCGC-1545/2020\|EPI_ISL_617966\|2020 | ···················· | ···················· | ···················· | ···················· | ···················· | ···················· |
| P | 2911 hCoV-19/Denmark/DCGC-1483/2020\|EPI_ISL_617967\|2020 | ···················· | ···················· | ···················· | ···················· | ···················· | ···················· |
| P | 2912 hCoV-19/Denmark/DCGC-1484/2020\|EPI_ISL_617968\|2020 | ···················· | ···················· | ···················· | ···················· | ···················· | ···················· |
| P | 2913 hCoV-19/Denmark/DCGC-1486/2020\|EPI_ISL_617970\|2020 | ···················· | ···················· | ···················· | ···················· | ···················· | ···················· |
| P | 2914 hCoV-19/Denmark/DCGC-1487/2020\|EPI_ISL_617971\|2020 | ···················· | ···················· | ···················· | ···················· | ···················· | ···················· |
| P | 2915 hCoV-19/Denmark/DCGC-1488/2020\|EPI_ISL_617972\|2020 | ···················· | ···················· | ···················· | ···················· | ···················· | ···················· |
| P | 2916 hCoV-19/Denmark/DCGC-1489/2020\|EPI_ISL_617973\|2020 | ···················· | ···················· | ···················· | ···················· | ···················· | ···················· |

| | | | | | | | | |
|---|---|---|---|---|---|---|---|---|
| P | 2917 | hCoV-19/Denmark/DCGC-1490/2020\|EPI_ISL_617974\|2020 | ···················· | ···················· | ····················· | ···················· | ···················· | ···················· |
| P | 2918 | hCoV-19/Denmark/DCGC-1492/2020\|EPI_ISL_617976\|2020 | ···················· | ···················· | ····················· | ···················· | ···················· | ···················· |
| P | 2919 | hCoV-19/Denmark/DCGC-1493/2020\|EPI_ISL_617977\|2020 | ···················· | ···················· | ····················· | ···················· | ···················· | ···················· |
| P | 2920 | hCoV-19/Denmark/DCGC-1495/2020\|EPI_ISL_617979\|2020 | ···················· | ···················· | ····················· | ···················· | ···················· | ···················· |
| P | 2921 | hCoV-19/Denmark/DCGC-1497/2020\|EPI_ISL_617981\|2020 | ···················· | ···················· | ····················· | ···················· | ···················· | ···················· |
| P | 2922 | hCoV-19/Denmark/DCGC-1498/2020\|EPI_ISL_617982\|2020 | ···················· | ···················· | ····················· | ···················· | ···················· | ···················· |
| P | 2923 | hCoV-19/Denmark/DCGC-1499/2020\|EPI_ISL_617983\|2020 | ···················· | ···················· | ····················· | ···················· | ···················· | ···················· |
| P | 2924 | hCoV-19/Denmark/DCGC-1500/2020\|EPI_ISL_617984\|2020 | ···················· | ···················· | ····················· | ···················· | ···················· | ···················· |
| P | 2925 | hCoV-19/Denmark/DCGC-1533/2020\|EPI_ISL_617985\|2020 | ···················· | ···················· | ····················· | ···················· | ···················· | ···················· |
| P | 2926 | hCoV-19/Denmark/DCGC-1534/2020\|EPI_ISL_617986\|2020 | ···················· | ···················· | ····················· | ···················· | ···················· | ···················· |
| P | 2927 | hCoV-19/Denmark/DCGC-1535/2020\|EPI_ISL_617987\|2020 | ···················· | ···················· | ····················· | ···················· | ···················· | ···················· |
| P | 2928 | hCoV-19/Denmark/DCGC-1537/2020\|EPI_ISL_617989\|2020 | ···················· | ···················· | ····················· | ···················· | ···················· | ···················· |
| P | 2929 | hCoV-19/Denmark/DCGC-6136/2020\|EPI_ISL_621170\|2020 | ···················· | ···················· | ····················· | ···················· | ···················· | ···················· |
| P | 2930 | hCoV-19/Denmark/DCGC-6137/2020\|EPI_ISL_621171\|2020 | ···················· | ···················· | ····················· | ···················· | ···················· | ···················· |
| P | 2931 | hCoV-19/Denmark/DCGC-6138/2020\|EPI_ISL_621172\|2020 | ···················· | ···················· | ····················· | ···················· | ···················· | ···················· |
| P | 2932 | hCoV-19/Denmark/DCGC-6139/2020\|EPI_ISL_621173\|2020 | ···················· | ···················· | ····················· | ···················· | ···················· | ···················· |
| P | 2933 | hCoV-19/Denmark/DCGC-6140/2020\|EPI_ISL_621174\|2020 | ···················· | ···················· | ····················· | ···················· | ···················· | ···················· |
| P | 2934 | hCoV-19/Denmark/DCGC-6143/2020\|EPI_ISL_621177\|2020 | ···················· | ···················· | ····················· | ···················· | ···················· | ···················· |
| P | 2935 | hCoV-19/Denmark/DCGC-6144/2020\|EPI_ISL_621178\|2020 | ···················· | ···················· | ····················· | ···················· | ···················· | ···················· |
| P | 2936 | hCoV-19/Denmark/DCGC-6145/2020\|EPI_ISL_621179\|2020 | ···················· | ···················· | ····················· | ···················· | ···················· | ···················· |
| P | 2937 | hCoV-19/Denmark/DCGC-6146/2020\|EPI_ISL_621180\|2020 | ···················· | ···················· | ····················· | ···················· | ···················· | ···················· |
| P | 2938 | hCoV-19/Denmark/DCGC-6147/2020\|EPI_ISL_621181\|2020 | ···················· | ···················· | ····················· | ···················· | ···················· | ···················· |
| P | 2939 | hCoV-19/Denmark/DCGC-6148/2020\|EPI_ISL_621182\|2020 | ···················· | ···················· | ····················· | ···················· | ···················· | ···················· |
| P | 2940 | hCoV-19/Denmark/DCGC-6149/2020\|EPI_ISL_621183\|2020 | ···················· | ···················· | ····················· | ···················· | ···················· | ···················· |
| P | 2941 | hCoV-19/Denmark/DCGC-6150/2020\|EPI_ISL_621184\|2020 | ···················· | ···················· | ····················· | ···················· | ···················· | ···················· |
| P | 2942 | hCoV-19/Denmark/DCGC-6151/2020\|EPI_ISL_621185\|2020 | ···················· | ···················· | ····················· | ···················· | ···················· | ···················· |
| P | 2943 | hCoV-19/Denmark/DCGC-6152/2020\|EPI_ISL_621186\|2020 | ···················· | ···················· | ····················· | ···················· | ···················· | ···················· |
| P | 2944 | hCoV-19/Denmark/DCGC-6330/2020\|EPI_ISL_621187\|2020 | ···················· | ···················· | ····················· | ···················· | ···················· | ···················· |
| P | 2945 | hCoV-19/Denmark/DCGC-5700/2020\|EPI_ISL_621205\|2020 | ···················· | ···················· | ····················· | ···················· | ···················· | ···················· |
| P | 2946 | hCoV-19/Denmark/DCGC-5701/2020\|EPI_ISL_621206\|2020 | ···················· | ···················· | ····················· | ···················· | ······T············· | ···················· |
| P | 2947 | hCoV-19/Denmark/DCGC-5702/2020\|EPI_ISL_621207\|2020 | ···················· | ···················· | ····················· | ···················· | ······T············· | ···················· |
| P | 2948 | hCoV-19/Denmark/DCGC-5703/2020\|EPI_ISL_621208\|2020 | ···················· | ···················· | ····················· | ···················· | ···················· | ···················· |
| P | 2949 | hCoV-19/Denmark/DCGC-5704/2020\|EPI_ISL_621209\|2020 | ···················· | ···················· | ····················· | ···················· | ···················· | ···················· |
| P | 2950 | hCoV-19/Denmark/DCGC-5705/2020\|EPI_ISL_621210\|2020 | ···················· | ···················· | ····················· | ···················· | ···················· | ···················· |
| P | 2951 | hCoV-19/Denmark/DCGC-5706/2020\|EPI_ISL_621211\|2020 | ···················· | ···················· | ····················· | ···················· | ···················· | ···················· |
| P | 2952 | hCoV-19/Denmark/DCGC-5707/2020\|EPI_ISL_621212\|2020 | ···················· | ···················· | ····················· | ···················· | ···················· | ···················· |
| P | 2953 | hCoV-19/Denmark/DCGC-5708/2020\|EPI_ISL_621213\|2020 | ···················· | ···················· | ····················· | ···················· | ···················· | ···················· |
| P | 2954 | hCoV-19/Denmark/DCGC-5709/2020\|EPI_ISL_621214\|2020 | ···················· | ···················· | ····················· | ···················· | ···················· | ···················· |
| P | 2955 | hCoV-19/Denmark/DCGC-5710/2020\|EPI_ISL_621215\|2020 | ···················· | ···················· | ····················· | ···················· | ···················· | ···················· |
| P | 2956 | hCoV-19/Denmark/DCGC-5711/2020\|EPI_ISL_621216\|2020 | ···················· | ···················· | ····················· | ···················· | ···················· | ···················· |
| P | 2957 | hCoV-19/Denmark/DCGC-5712/2020\|EPI_ISL_621217\|2020 | ···················· | ···················· | ····················· | ···················· | ···················· | ···················· |
| P | 2958 | hCoV-19/Denmark/DCGC-5713/2020\|EPI_ISL_621218\|2020 | ···················· | ···················· | ····················· | ···················· | ···················· | ···················· |
| P | 2959 | hCoV-19/Denmark/DCGC-5714/2020\|EPI_ISL_621219\|2020 | ···················· | ···················· | ····················· | ···················· | ···················· | ···················· |
| P | 2960 | hCoV-19/Denmark/DCGC-5715/2020\|EPI_ISL_621220\|2020 | ···················· | ···················· | ····················· | ···················· | ···················· | ···················· |
| P | 2961 | hCoV-19/Denmark/DCGC-5716/2020\|EPI_ISL_621221\|2020 | ···················· | ···················· | ····················· | ···················· | ···················· | ···················· |
| P | 2962 | hCoV-19/Denmark/DCGC-5717/2020\|EPI_ISL_621222\|2020 | ···················· | ···················· | ····················· | ···················· | ···················· | ···················· |
| P | 2963 | hCoV-19/Denmark/DCGC-5718/2020\|EPI_ISL_621223\|2020 | ···················· | ···················· | ····················· | ···················· | ···················· | ···················· |
| P | 2964 | hCoV-19/Denmark/DCGC-5719/2020\|EPI_ISL_621224\|2020 | ···················· | ···················· | ····················· | ···················· | ···················· | ···················· |
| P | 2965 | hCoV-19/Denmark/DCGC-5720/2020\|EPI_ISL_621225\|2020 | ···················· | ···················· | ····················· | ···················· | ···················· | ···················· |
| P | 2966 | hCoV-19/Denmark/DCGC-5721/2020\|EPI_ISL_621226\|2020 | ···················· | ···················· | ····················· | ···················· | ···················· | ···················· |
| P | 2967 | hCoV-19/Denmark/DCGC-5722/2020\|EPI_ISL_621227\|2020 | ···················· | ···················· | ····················· | ···················· | ···················· | ···················· |
| P | 2968 | hCoV-19/Denmark/DCGC-5723/2020\|EPI_ISL_621228\|2020 | ···················· | ···················· | ····················· | ···················· | ···················· | ···················· |
| P | 2969 | hCoV-19/Denmark/DCGC-5724/2020\|EPI_ISL_621229\|2020 | ···················· | ···················· | ····················· | ···················· | ···················· | ···················· |
| P | 2970 | hCoV-19/Denmark/DCGC-5725/2020\|EPI_ISL_621230\|2020 | ···················· | ···················· | ····················· | ···················· | ···················· | ···················· |
| P | 2971 | hCoV-19/Denmark/DCGC-5726/2020\|EPI_ISL_621231\|2020 | ···················· | ···················· | ····················· | ···················· | ···················· | ···················· |
| P | 2972 | hCoV-19/Denmark/DCGC-5727/2020\|EPI_ISL_621232\|2020 | ···················· | ···················· | ····················· | ···················· | ···················· | ···················· |
| P | 2973 | hCoV-19/Denmark/DCGC-5728/2020\|EPI_ISL_621233\|2020 | ···················· | ···················· | ····················· | ···················· | ···················· | ···················· |
| P | 2974 | hCoV-19/Denmark/DCGC-5729/2020\|EPI_ISL_621234\|2020 | ···················· | ···················· | ····················· | ···················· | ···················· | ···················· |
| P | 2975 | hCoV-19/Denmark/DCGC-5730/2020\|EPI_ISL_621235\|2020 | ···················· | ···················· | ····················· | ···················· | ···················· | ···················· |
| P | 2976 | hCoV-19/Denmark/DCGC-5731/2020\|EPI_ISL_621236\|2020 | ···················· | ···················· | ····················· | ···················· | ···················· | ···················· |
| P | 2977 | hCoV-19/Denmark/DCGC-5732/2020\|EPI_ISL_621237\|2020 | ···················· | ···················· | ····················· | ···················· | ···················· | ···················· |
| P | 2978 | hCoV-19/Denmark/DCGC-5733/2020\|EPI_ISL_621238\|2020 | ···················· | ···················· | ····················· | ···················· | ···················· | ···················· |
| P | 2979 | hCoV-19/Denmark/DCGC-5734/2020\|EPI_ISL_621239\|2020 | ···················· | ···················· | ····················· | ···················· | ···················· | ···················· |
| P | 2980 | hCoV-19/Denmark/DCGC-5735/2020\|EPI_ISL_621240\|2020 | ···················· | ···················· | ····················· | ···················· | ···················· | ···················· |
| P | 2981 | hCoV-19/Denmark/DCGC-5736/2020\|EPI_ISL_621241\|2020 | ···················· | ···················· | ····················· | ···················· | ······T············· | ···················· |
| P | 2982 | hCoV-19/Denmark/DCGC-5737/2020\|EPI_ISL_621242\|2020 | ···················· | ···················· | ····················· | ···················· | ···················· | ···················· |
| P | 2983 | hCoV-19/Denmark/DCGC-5738/2020\|EPI_ISL_621243\|2020 | ···················· | ···················· | ····················· | ···················· | ···················· | ···················· |
| P | 2984 | hCoV-19/Denmark/DCGC-5739/2020\|EPI_ISL_621244\|2020 | ···················· | ···················· | ····················· | ···················· | ···················· | ···················· |
| P | 2985 | hCoV-19/Denmark/DCGC-5740/2020\|EPI_ISL_621245\|2020 | ···················· | ···················· | ····················· | ···················· | ···················· | ···················· |
| P | 2986 | hCoV-19/Denmark/DCGC-5741/2020\|EPI_ISL_621246\|2020 | ···················· | ···················· | ····················· | ···················· | ···················· | ···················· |
| P | 2987 | hCoV-19/Denmark/DCGC-5742/2020\|EPI_ISL_621247\|2020 | ···················· | ···················· | ····················· | ···················· | ···················· | ···················· |
| P | 2988 | hCoV-19/Denmark/DCGC-5743/2020\|EPI_ISL_621248\|2020 | ···················· | ···················· | ····················· | ···················· | ···················· | ···················· |
| P | 2989 | hCoV-19/Denmark/DCGC-4601/2020\|EPI_ISL_621270\|2020 | ···················· | ···················· | ····················· | ···················· | ···················· | ···················· |
| P | 2990 | hCoV-19/Denmark/DCGC-4602/2020\|EPI_ISL_621271\|2020 | ···················· | ···················· | ····················· | ···················· | ···················· | ···················· |
| P | 2991 | hCoV-19/Denmark/DCGC-4603/2020\|EPI_ISL_621272\|2020 | ···················· | ···················· | ····················· | ···················· | ···················· | ···················· |
| P | 2992 | hCoV-19/Denmark/DCGC-4604/2020\|EPI_ISL_621273\|2020 | ···················· | ···················· | ····················· | ···················· | ···················· | ···················· |
| P | 2993 | hCoV-19/Denmark/DCGC-4605/2020\|EPI_ISL_621274\|2020 | ···················· | ···················· | ····················· | ···················· | ···················· | ···················· |
| P | 2994 | hCoV-19/Denmark/DCGC-4606/2020\|EPI_ISL_621275\|2020 | ···················· | ···················· | ····················· | ···················· | ···················· | ···················· |
| P | 2995 | hCoV-19/Denmark/DCGC-4607/2020\|EPI_ISL_621276\|2020 | ···················· | ···················· | ····················· | ···················· | ···················· | ···················· |
| P | 2996 | hCoV-19/Denmark/DCGC-4608/2020\|EPI_ISL_621277\|2020 | ···················· | ···················· | ····················· | ···················· | ···················· | ···················· |

P 2997 hCoV-19/Denmark/DCGC-4609/2020|EPI_ISL_621278|2020 ···················· ···················· ···················· ···················· ···················· ····················
P 2998 hCoV-19/Denmark/DCGC-4610/2020|EPI_ISL_621279|2020 ···················· ···················· ···················· ···················· ···················· ····················
P 2999 hCoV-19/Denmark/DCGC-4618/2020|EPI_ISL_621287|2020 ···················· ···················· ···················· ···················· ···················· ····················
P 3000 hCoV-19/Denmark/DCGC-4619/2020|EPI_ISL_621288|2020 ···················· ···················· ···················· ···················· ···················· ····················
P 3001 hCoV-19/Denmark/DCGC-4620/2020|EPI_ISL_621289|2020 ···················· ···················· ···················· ···················· ···················· ····················
P 3002 hCoV-19/Denmark/DCGC-1848/2020|EPI_ISL_622536|2020 ···················· ···················· ···················· ···················· ···················· ····················
P 3003 hCoV-19/Denmark/DCGC-1853/2020|EPI_ISL_622537|2020 ···················· ···················· ···················· ···················· ···················· ····················
P 3004 hCoV-19/Denmark/DCGC-1867/2020|EPI_ISL_622539|2020 ········T··········· ···················· ··········T········· ···················· ···················· ····················
P 3005 hCoV-19/Denmark/DCGC-1875/2020|EPI_ISL_622540|2020 ···················· ···················· ···················· ···················· ···················· ····················
P 3006 hCoV-19/Denmark/DCGC-1881/2020|EPI_ISL_622541|2020 ···················· ···················· ···················· ···················· ···················· ····················
P 3007 hCoV-19/Denmark/DCGC-1886/2020|EPI_ISL_622542|2020 ···················· ···················· ···················· ···················· ···················· ····················
P 3008 hCoV-19/Denmark/DCGC-1894/2020|EPI_ISL_622543|2020 ···················· ···················· ···················· ···················· ···················· ····················
P 3009 hCoV-19/Denmark/DCGC-1900/2020|EPI_ISL_622544|2020 ···················· ···················· ···················· ···················· ···················· ····················
P 3010 hCoV-19/Denmark/DCGC-1826/2020|EPI_ISL_622545|2020 ···················· ···················· ···················· ···················· ···················· ····················
P 3011 hCoV-19/Denmark/DCGC-1834/2020|EPI_ISL_622546|2020 ···················· ···················· ···················· ···················· ···················· ····················
P 3012 hCoV-19/Denmark/DCGC-1854/2020|EPI_ISL_622549|2020 ···················· ···················· ···················· ···················· ···················· ····················
P 3013 hCoV-19/Denmark/DCGC-1868/2020|EPI_ISL_622550|2020 ···················· ···················· ···················· ···················· ···················· ····················
P 3014 hCoV-19/Denmark/DCGC-1887/2020|EPI_ISL_622551|2020 ···················· ···················· ···················· ···················· ···················· ····················
P 3015 hCoV-19/Denmark/DCGC-1895/2020|EPI_ISL_622552|2020 ···················· ···················· ···················· ···················· ···················· ····················
P 3016 hCoV-19/Denmark/DCGC-1827/2020|EPI_ISL_622553|2020 ···················· ···················· ···················· ···················· ···················· ····················
P 3017 hCoV-19/Denmark/DCGC-1835/2020|EPI_ISL_622554|2020 ···················· ···················· ···················· ···················· ···················· ····················
P 3018 hCoV-19/Denmark/DCGC-1843/2020|EPI_ISL_622555|2020 ···················· ···················· ···················· ···················· ···················· ····················
P 3019 hCoV-19/Denmark/DCGC-1937/2020|EPI_ISL_622556|2020 ···················· ···················· ···················· ···················· ···················· ····················
P 3020 hCoV-19/Denmark/DCGC-1855/2020|EPI_ISL_622557|2020 ···················· ···················· ···················· ···················· ···················· ····················
P 3021 hCoV-19/Denmark/DCGC-1862/2020|EPI_ISL_622558|2020 ···················· ···················· ···················· ···················· ···················· ····················
P 3022 hCoV-19/Denmark/DCGC-1869/2020|EPI_ISL_622559|2020 ···················· ···················· ···················· ···················· ···················· ····················
P 3023 hCoV-19/Denmark/DCGC-1876/2020|EPI_ISL_622560|2020 ···················· ···················· ···················· ···················· ···················· ····················
P 3024 hCoV-19/Denmark/DCGC-1939/2020|EPI_ISL_622561|2020 ···················· ···················· ···················· ···················· ···················· ····················
P 3025 hCoV-19/Denmark/DCGC-1888/2020|EPI_ISL_622562|2020 ···················· ···················· ···················· ···················· ···················· ····················
P 3026 hCoV-19/Denmark/DCGC-1896/2020|EPI_ISL_622563|2020 ···················· ···················· ···················· ···················· ···················· ····················
P 3027 hCoV-19/Denmark/DCGC-1901/2020|EPI_ISL_622564|2020 ···················· ···················· ···················· ···················· ···················· ····················
P 3028 hCoV-19/Denmark/DCGC-1828/2020|EPI_ISL_622565|2020 ···················· ···················· ···················· ···················· ···················· ····················
P 3029 hCoV-19/Denmark/DCGC-1836/2020|EPI_ISL_622566|2020 ···················· ···················· ···················· ···················· ···················· ····················
P 3030 hCoV-19/Denmark/DCGC-1844/2020|EPI_ISL_622567|2020 ···················· ···················· ···················· ···················· ···················· ····················
P 3031 hCoV-19/Denmark/DCGC-1850/2020|EPI_ISL_622568|2020 ···················· ···················· ···················· ···················· ···················· ····················
P 3032 hCoV-19/Denmark/DCGC-1856/2020|EPI_ISL_622569|2020 ···················· ···················· ···················· ···················· ···················· ····················
P 3033 hCoV-19/Denmark/DCGC-1863/2020|EPI_ISL_622570|2020 ···················· ···················· ···················· ···················· ···················· ····················
P 3034 hCoV-19/Denmark/DCGC-1870/2020|EPI_ISL_622571|2020 ···················· ···················· ···················· ···················· ···················· ····················
P 3035 hCoV-19/Denmark/DCGC-1877/2020|EPI_ISL_622572|2020 ···················· ···················· ···················· ···················· ···················· ····················
P 3036 hCoV-19/Denmark/DCGC-1882/2020|EPI_ISL_622573|2020 ···················· ···················· ···················· ···················· ···················· ····················
P 3037 hCoV-19/Denmark/DCGC-1889/2020|EPI_ISL_622574|2020 ···················· ···················· ···················· ···················· ···················· ····················
P 3038 hCoV-19/Denmark/DCGC-1829/2020|EPI_ISL_622575|2020 ···················· ···················· ···················· ···················· ···················· ····················
P 3039 hCoV-19/Denmark/DCGC-1837/2020|EPI_ISL_622576|2020 ···················· ···················· ···················· ···················· ···················· ····················
P 3040 hCoV-19/Denmark/DCGC-1851/2020|EPI_ISL_622577|2020 ········T··········· ···················· ··········T········· ···················· ···················· ····················
P 3041 hCoV-19/Denmark/DCGC-1857/2020|EPI_ISL_622578|2020 ···················· ···················· ···················· ···················· ···················· ····················
P 3042 hCoV-19/Denmark/DCGC-1864/2020|EPI_ISL_622579|2020 ···················· ···················· ···················· ···················· ···················· ····················
P 3043 hCoV-19/England/ALDP-AA3120/2020|EPI_ISL_623867|20 ···················· ···················· ···················· ···················· ···················· ····················
P 3044 hCoV-19/England/QEUH-AAEF76/2020|EPI_ISL_623868|20 ···················· ···················· ···················· ···················· ···················· ····················
P 3045 hCoV-19/England/ALDP-AA0D98/2020|EPI_ISL_623869|20 ···················· ···················· ···················· ···················· ···················· ···A················
P 3046 hCoV-19/England/ALDP-AA2EFD/2020|EPI_ISL_623870|20 ···················· ···················· ···················· ···················· ···················· ····················
P 3047 hCoV-19/England/ALDP-AA51E2/2020|EPI_ISL_623871|20 ···················· ···················· ···················· ···················· ···················· ····················
P 3048 hCoV-19/England/QEUH-AAE495/2020|EPI_ISL_623873|20 ···················· ···················· ···················· ···················· ···················· ····················
P 3049 hCoV-19/England/ALDP-AA14A1/2020|EPI_ISL_623875|20 ···················· ···················· ···················· ···················· ···················· ····················
P 3050 hCoV-19/Scotland/QEUH-AADD40/2020|EPI_ISL_623876|2 ···················· ···················· ···················· ···················· ···················· ····················
P 3051 hCoV-19/England/ALDP-AA522B/2020|EPI_ISL_623877|20 ···················· ···················· ···················· ···················· ···················· ····················
P 3052 hCoV-19/England/ALDP-AA41A7/2020|EPI_ISL_623879|20 ···················· ···················· ···················· ···················· ···················· ····················
P 3053 hCoV-19/England/ALDP-AA34DC/2020|EPI_ISL_623880|20 ···················· ···················· ···················· ···················· ···················· ····················
P 3054 hCoV-19/England/ALDP-AA3102/2020|EPI_ISL_623881|20 ···················· ···················· ···················· ···················· ···················· ····················
P 3055 hCoV-19/England/ALDP-AA2BBA/2020|EPI_ISL_623882|20 ···················· ···················· ···················· ···················· ···················· ····················
P 3056 hCoV-19/Scotland/QEUH-AAEF0D/2020|EPI_ISL_623883|2 ···················· ···················· ···················· ···················· ···················· ····················
P 3057 hCoV-19/England/QEUH-AAE565/2020|EPI_ISL_623884|20 ···················· ···················· ···················· ···················· ···················· ····················
P 3058 hCoV-19/England/ALDP-AA523A/2020|EPI_ISL_623885|20 ···················· ···················· ···················· ···················· ···················· ····················
P 3059 hCoV-19/England/QEUH-AADC9E/2020|EPI_ISL_623886|20 ···················· ···················· ···················· ···················· ···················· ····················
P 3060 hCoV-19/England/ALDP-AA16F6/2020|EPI_ISL_623887|20 ···················· ···················· ···················· ···················· ···················· ····················
P 3061 hCoV-19/England/ALDP-AA133B/2020|EPI_ISL_623888|20 ···················· ···················· ···················· ···················· ···················· ····················
P 3062 hCoV-19/England/ALDP-AA07A9/2020|EPI_ISL_623889|20 ···················· ···················· ···················· ···················· ···················· ····················
P 3063 hCoV-19/England/ALDP-AA34CD/2020|EPI_ISL_623890|20 ···················· ···················· ···················· ···················· ···················· ····················
P 3064 hCoV-19/England/ALDP-AA1456/2020|EPI_ISL_623891|20 ···················· ···················· ···················· ···················· ···················· ····················
P 3065 hCoV-19/England/ALDP-AA5698/2020|EPI_ISL_623892|20 ···················· ···················· ···················· ···················· ···················· ····················
P 3066 hCoV-19/England/QEUH-AADC07/2020|EPI_ISL_623893|20 ···················· ···················· ···················· ···················· ···················· ····················
P 3067 hCoV-19/England/ALDP-AA2DA5/2020|EPI_ISL_623894|20 ···················· ···················· ···················· ···················· ······T············· ····················
P 3068 hCoV-19/England/QEUH-AAD3A8/2020|EPI_ISL_623895|20 ···················· ···················· ···················· ···················· ···················· ····················
P 3069 hCoV-19/England/ALDP-AA5267/2020|EPI_ISL_623896|20 ···················· ···················· ···················· ···················· ···················· ····················
P 3070 hCoV-19/England/ALDP-AA1438/2020|EPI_ISL_623897|20 ···················· ···················· ···················· ···················· ···················· ····················
P 3071 hCoV-19/England/ALDP-AA13B3/2020|EPI_ISL_623898|20 ···················· ···················· ···················· ···················· ···················· ····················
P 3072 hCoV-19/England/ALDP-AA1447/2020|EPI_ISL_623899|20 ············T······· ···················· ···················· ···················· ···················· ····················
P 3073 hCoV-19/Denmark/DCGC-5398/2020|EPI_ISL_615330|2020 ···················· ···················· ···················· ···················· ···················· ····················
P 3074 hCoV-19/Denmark/DCGC-5399/2020|EPI_ISL_615331|2020 ···················· ···················· ···················· ···················· ···················· ····················
P 3075 hCoV-19/Denmark/DCGC-5400/2020|EPI_ISL_615332|2020 ···················· ···················· ···················· ···················· ···················· ····················
P 3076 hCoV-19/Denmark/DCGC-5401/2020|EPI_ISL_615333|2020 ···················· ···················· ···················· ···················· ···················· ····················

| | | | | | | | |
|---|---|---|---|---|---|---|---|
| P | 3077 hCoV-19/Denmark/DCGC-5402/2020\|EPI_ISL_615334\|2020 | .................... | .................... | ..................... | .................... | .................... | .................... |
| P | 3078 hCoV-19/Denmark/DCGC-5404/2020\|EPI_ISL_615336\|2020 | .................... | .................... | ..................... | .................... | .................... | .................... |
| P | 3079 hCoV-19/Denmark/DCGC-5405/2020\|EPI_ISL_615337\|2020 | .................... | .................... | ..................... | .................... | .................... | .................... |
| P | 3080 hCoV-19/Denmark/DCGC-5406/2020\|EPI_ISL_615338\|2020 | .................... | .................... | ..................... | .................... | .................... | .................... |
| P | 3081 hCoV-19/Denmark/DCGC-5407/2020\|EPI_ISL_615339\|2020 | .................... | .................... | ..................... | .................... | .................... | .................... |
| P | 3082 hCoV-19/Denmark/DCGC-5408/2020\|EPI_ISL_615340\|2020 | .................... | .................... | ..................... | .................... | .................... | .................... |
| P | 3083 hCoV-19/Denmark/DCGC-5409/2020\|EPI_ISL_615341\|2020 | .................... | .................... | ..................... | .................... | .................... | .................... |
| P | 3084 hCoV-19/Denmark/DCGC-5410/2020\|EPI_ISL_615342\|2020 | .................... | .................... | ..................... | .................... | .................... | .................... |
| P | 3085 hCoV-19/Denmark/DCGC-5411/2020\|EPI_ISL_615343\|2020 | .................... | .................... | ..................... | .................... | .................... | .................... |
| P | 3086 hCoV-19/Denmark/DCGC-5412/2020\|EPI_ISL_615344\|2020 | .................... | .................... | ..................... | .................... | .................... | .................... |
| P | 3087 hCoV-19/Denmark/DCGC-5414/2020\|EPI_ISL_615346\|2020 | .................... | .................... | ..................... | .................... | .................... | .................... |
| P | 3088 hCoV-19/Denmark/DCGC-5415/2020\|EPI_ISL_615347\|2020 | .................... | .................... | ..................... | .................... | .................... | .................... |
| P | 3089 hCoV-19/Denmark/DCGC-5416/2020\|EPI_ISL_615348\|2020 | .................... | .................... | ..................... | .................... | .................... | .................... |
| P | 3090 hCoV-19/Denmark/DCGC-5417/2020\|EPI_ISL_615349\|2020 | .................... | .................... | ..................... | .................... | .................... | .................... |
| P | 3091 hCoV-19/Denmark/DCGC-5418/2020\|EPI_ISL_615350\|2020 | .................... | .................... | ..................... | .................... | .................... | .................... |
| P | 3092 hCoV-19/Denmark/DCGC-5419/2020\|EPI_ISL_615351\|2020 | .................... | .................... | ..................... | .................... | .................... | .................... |
| P | 3093 hCoV-19/Denmark/DCGC-5420/2020\|EPI_ISL_615352\|2020 | .................... | .................... | ..................... | .................... | .................... | .................... |
| P | 3094 hCoV-19/Denmark/DCGC-5421/2020\|EPI_ISL_615353\|2020 | .................... | .................... | ..................... | .................... | .................... | .................... |
| P | 3095 hCoV-19/Denmark/DCGC-5422/2020\|EPI_ISL_615354\|2020 | .................... | .................... | ..................... | .................... | .................... | .................... |
| P | 3096 hCoV-19/Denmark/DCGC-5423/2020\|EPI_ISL_615355\|2020 | .................... | .................... | ..................... | .................... | .................... | .................... |
| P | 3097 hCoV-19/Denmark/DCGC-5424/2020\|EPI_ISL_615356\|2020 | .................... | .................... | ..................... | .................... | .................... | .................... |
| P | 3098 hCoV-19/Denmark/DCGC-5425/2020\|EPI_ISL_615357\|2020 | .................... | .................... | ..................... | .................... | .................... | .................... |
| P | 3099 hCoV-19/Denmark/DCGC-5426/2020\|EPI_ISL_615358\|2020 | .................... | .................... | ..................... | .................... | .................... | .................... |
| P | 3100 hCoV-19/Denmark/DCGC-5427/2020\|EPI_ISL_615359\|2020 | .................... | .................... | ..................... | .................... | .................... | .................... |
| P | 3101 hCoV-19/Denmark/DCGC-5429/2020\|EPI_ISL_615361\|2020 | .................... | .................... | ..................... | .................... | .................... | .................... |
| P | 3102 hCoV-19/Denmark/DCGC-5430/2020\|EPI_ISL_615362\|2020 | .................... | .................... | ..................... | .................... | .................... | .................... |
| P | 3103 hCoV-19/Denmark/DCGC-5431/2020\|EPI_ISL_615363\|2020 | .................... | .................... | ..................... | .................... | .................... | .................... |
| P | 3104 hCoV-19/Denmark/DCGC-5432/2020\|EPI_ISL_615364\|2020 | .................... | .................... | ..................... | .................... | .................... | .................... |
| P | 3105 hCoV-19/Denmark/DCGC-5433/2020\|EPI_ISL_615365\|2020 | .................... | .................... | ..................... | .................... | .................... | .................... |
| P | 3106 hCoV-19/Denmark/DCGC-5434/2020\|EPI_ISL_615366\|2020 | .................... | .................... | ..................... | .................... | .................... | .................... |
| P | 3107 hCoV-19/Denmark/DCGC-5435/2020\|EPI_ISL_615367\|2020 | .................... | .................... | ..................... | .................... | .................... | .................... |
| P | 3108 hCoV-19/Denmark/DCGC-5436/2020\|EPI_ISL_615368\|2020 | .................... | .................... | ..................... | .................... | .................... | .................... |
| P | 3109 hCoV-19/Denmark/DCGC-5437/2020\|EPI_ISL_615369\|2020 | .................... | .................... | ..................... | .................... | .................... | .................... |
| P | 3110 hCoV-19/Denmark/DCGC-4509/2020\|EPI_ISL_615370\|2020 | .................... | .................... | ..................... | .................... | .................... | .................... |
| P | 3111 hCoV-19/Denmark/DCGC-4510/2020\|EPI_ISL_615371\|2020 | .................... | .................... | ..................... | .................... | .................... | .................... |
| P | 3112 hCoV-19/Denmark/DCGC-4511/2020\|EPI_ISL_615372\|2020 | .................... | .................... | ..................... | .................... | .................... | .................... |
| P | 3113 hCoV-19/Denmark/DCGC-4512/2020\|EPI_ISL_615373\|2020 | .................... | .................... | ..................... | .................... | .................... | .................... |
| P | 3114 hCoV-19/Denmark/DCGC-4513/2020\|EPI_ISL_615374\|2020 | .................... | .................... | ..................... | .................... | .................... | .................... |
| P | 3115 hCoV-19/Denmark/DCGC-4514/2020\|EPI_ISL_615375\|2020 | .................... | .................... | ..................... | .................... | .................... | .................... |
| P | 3116 hCoV-19/Denmark/DCGC-4378/2020\|EPI_ISL_615376\|2020 | .................... | .................... | ..................... | .................... | .................... | .................... |
| P | 3117 hCoV-19/Denmark/DCGC-4379/2020\|EPI_ISL_615377\|2020 | .................... | .................... | ..................... | .................... | .................... | .................... |
| P | 3118 hCoV-19/Denmark/DCGC-4380/2020\|EPI_ISL_615378\|2020 | .................... | .................... | ..................... | .................... | .................... | .................... |
| P | 3119 hCoV-19/Denmark/DCGC-4381/2020\|EPI_ISL_615379\|2020 | .................... | .................... | ..................... | .................... | .................... | .................... |
| P | 3120 hCoV-19/Denmark/DCGC-4382/2020\|EPI_ISL_615380\|2020 | .................... | .................... | ..................... | .................... | .................... | .................... |
| P | 3121 hCoV-19/Denmark/DCGC-4383/2020\|EPI_ISL_615381\|2020 | .................... | .................... | ..................... | .................... | .................... | .................... |
| P | 3122 hCoV-19/Denmark/DCGC-4384/2020\|EPI_ISL_615382\|2020 | .................... | .................... | ..................... | .................... | .................... | .................... |
| P | 3123 hCoV-19/Denmark/DCGC-4385/2020\|EPI_ISL_615383\|2020 | .................... | .................... | ..................... | .................... | .................... | .................... |
| P | 3124 hCoV-19/Denmark/DCGC-4386/2020\|EPI_ISL_615384\|2020 | .................... | .................... | ..................... | .................... | .................... | .................... |
| P | 3125 hCoV-19/Denmark/DCGC-4387/2020\|EPI_ISL_615385\|2020 | .................... | .................... | ..................... | .................... | .................... | .................... |
| P | 3126 hCoV-19/Denmark/DCGC-4388/2020\|EPI_ISL_615386\|2020 | .................... | .................... | ..................... | .................... | .................... | .................... |
| P | 3127 hCoV-19/Denmark/DCGC-4389/2020\|EPI_ISL_615387\|2020 | .................... | .................... | ..................... | .................... | .................... | .................... |
| P | 3128 hCoV-19/Denmark/DCGC-4390/2020\|EPI_ISL_615388\|2020 | .................... | .................... | ..................... | .................... | .................... | .................... |
| P | 3129 hCoV-19/Denmark/DCGC-4391/2020\|EPI_ISL_615389\|2020 | .................... | .................... | ..................... | .................... | .................... | .................... |
| P | 3130 hCoV-19/Denmark/DCGC-4392/2020\|EPI_ISL_615390\|2020 | .................... | .................... | ..................... | .................... | .................... | .................... |
| P | 3131 hCoV-19/Denmark/DCGC-4393/2020\|EPI_ISL_615391\|2020 | .................... | .................... | ..................... | .................... | .................... | .................... |
| P | 3132 hCoV-19/Denmark/DCGC-4394/2020\|EPI_ISL_615392\|2020 | .................... | .................... | ..................... | .................... | .................... | .................... |
| P | 3133 hCoV-19/Denmark/DCGC-4396/2020\|EPI_ISL_615394\|2020 | .................... | .................... | ..................... | .................... | .................... | .................... |
| P | 3134 hCoV-19/Denmark/DCGC-4397/2020\|EPI_ISL_615395\|2020 | .................... | .................... | ..................... | .................... | .................... | .................... |
| P | 3135 hCoV-19/Denmark/DCGC-4398/2020\|EPI_ISL_615396\|2020 | .................... | .................... | ..................... | .................... | .................... | .................... |
| P | 3136 hCoV-19/Denmark/DCGC-4400/2020\|EPI_ISL_615398\|2020 | .................... | .................... | ..................... | .................... | .................... | .................... |
| P | 3137 hCoV-19/Denmark/DCGC-4401/2020\|EPI_ISL_615399\|2020 | .................... | .................... | ..................... | .................... | .................... | .................... |
| P | 3138 hCoV-19/Denmark/DCGC-2766/2020\|EPI_ISL_616600\|2020 | .................... | .................... | ..................... | .................... | .................... | .................... |
| P | 3139 hCoV-19/Denmark/DCGC-2767/2020\|EPI_ISL_616601\|2020 | .................... | .................... | ..................... | .................... | .................... | .................... |
| P | 3140 hCoV-19/Denmark/DCGC-2768/2020\|EPI_ISL_616602\|2020 | .................... | .................... | ..................... | .................... | .................... | .................... |
| P | 3141 hCoV-19/Denmark/DCGC-2769/2020\|EPI_ISL_616603\|2020 | .................... | .................... | ..................... | .................... | .................... | .................... |
| P | 3142 hCoV-19/Denmark/DCGC-2770/2020\|EPI_ISL_616604\|2020 | .................... | .................... | ..................... | .................... | .................... | .................... |
| P | 3143 hCoV-19/Denmark/DCGC-2771/2020\|EPI_ISL_616605\|2020 | .................... | .................... | ..................... | .................... | .................... | .................... |
| P | 3144 hCoV-19/Denmark/DCGC-2925/2020\|EPI_ISL_616606\|2020 | .................... | .................... | ..................... | .................... | .................... | .................... |
| P | 3145 hCoV-19/Denmark/DCGC-2772/2020\|EPI_ISL_616607\|2020 | .................... | .................... | ..................... | .................... | .................... | .................... |
| P | 3146 hCoV-19/Denmark/DCGC-2773/2020\|EPI_ISL_616608\|2020 | .................... | .................... | ..................... | .................... | .................... | .................... |
| P | 3147 hCoV-19/Denmark/DCGC-2774/2020\|EPI_ISL_616609\|2020 | .................... | ......C............. | ..................... | .................... | .................... | .................... |
| P | 3148 hCoV-19/Denmark/DCGC-2775/2020\|EPI_ISL_616610\|2020 | .................... | .................... | ..................... | .................... | .................... | .................... |
| P | 3149 hCoV-19/Denmark/DCGC-2776/2020\|EPI_ISL_616611\|2020 | .................... | .................... | ..................... | .................... | .................... | .................... |
| P | 3150 hCoV-19/Denmark/DCGC-2777/2020\|EPI_ISL_616612\|2020 | .................... | .................... | ..................... | .................... | .................... | .................... |
| P | 3151 hCoV-19/Denmark/DCGC-2778/2020\|EPI_ISL_616613\|2020 | .................... | .................... | ..................... | .................... | .................... | .................... |
| P | 3152 hCoV-19/Denmark/DCGC-2780/2020\|EPI_ISL_616615\|2020 | .................... | .................... | ..................... | .................... | .................... | .................... |
| P | 3153 hCoV-19/Denmark/DCGC-2781/2020\|EPI_ISL_616616\|2020 | .................... | .................... | ..................... | .................... | .................... | .................... |
| P | 3154 hCoV-19/Denmark/DCGC-2782/2020\|EPI_ISL_616617\|2020 | .................... | .................... | ..................... | .................... | .................... | .................... |
| P | 3155 hCoV-19/Denmark/DCGC-2834/2020\|EPI_ISL_616660\|2020 | .................... | .................... | ..................... | .................... | .................... | .................... |
| P | 3156 hCoV-19/Denmark/DCGC-2835/2020\|EPI_ISL_616661\|2020 | .................... | .................... | ..................... | .................... | .................... | .................... |

| | | | | | | | |
|---|---|---|---|---|---|---|---|
| P | 3157 hCoV-19/Denmark/DCGC-2928/2020\|EPI_ISL_616662\|2020 | ···················· | ···················· | ···················· | ···················· | ···················· | ···················· |
| P | 3158 hCoV-19/Denmark/DCGC-2836/2020\|EPI_ISL_616663\|2020 | ···················· | ···················· | ···················· | ···················· | ···················· | ···················· |
| P | 3159 hCoV-19/Denmark/DCGC-2837/2020\|EPI_ISL_616664\|2020 | ···················· | ···················· | ···················· | ···················· | ···················· | ···················· |
| P | 3161 hCoV-19/Denmark/DCGC-2839/2020\|EPI_ISL_616666\|2020 | ···················· | ···················· | ···················· | ···················· | ···················· | ···················· |
| P | 3162 hCoV-19/Denmark/DCGC-2840/2020\|EPI_ISL_616667\|2020 | ···················· | ···················· | ···················· | ···················· | ·····T·············· | ···················· |
| P | 3163 hCoV-19/Denmark/DCGC-2841/2020\|EPI_ISL_616668\|2020 | ···················· | ···················· | ···················· | ···················· | ···················· | ···················· |
| P | 3164 hCoV-19/Denmark/DCGC-2842/2020\|EPI_ISL_616669\|2020 | ···················· | ···················· | ···················· | ···················· | ···················· | ···················· |
| P | 3165 hCoV-19/Denmark/DCGC-2843/2020\|EPI_ISL_616670\|2020 | ···················· | ···················· | ···················· | ···················· | ···················· | ···················· |
| P | 3166 hCoV-19/Denmark/DCGC-2844/2020\|EPI_ISL_616671\|2020 | ···················· | ···················· | ···················· | ···················· | ···················· | ···················· |
| P | 3167 hCoV-19/Denmark/DCGC-2845/2020\|EPI_ISL_616672\|2020 | ···················· | ···················· | ···················· | ···················· | ···················· | ···················· |
| P | 3168 hCoV-19/Denmark/DCGC-2846/2020\|EPI_ISL_616673\|2020 | ···················· | ···················· | ···················· | ···················· | ···················· | ···················· |
| P | 3169 hCoV-19/Denmark/DCGC-2847/2020\|EPI_ISL_616674\|2020 | ···················· | ·A·················· | ···················· | ···················· | ···················· | ···················· |
| P | 3170 hCoV-19/Denmark/DCGC-2848/2020\|EPI_ISL_616675\|2020 | ···················· | ···················· | ···················· | ···················· | ···················· | ···················· |
| P | 3171 hCoV-19/Denmark/DCGC-2849/2020\|EPI_ISL_616676\|2020 | ···················· | ···················· | ···················· | ···················· | ···················· | ···················· |
| P | 3172 hCoV-19/Denmark/DCGC-2850/2020\|EPI_ISL_616677\|2020 | ···················· | ···················· | ···················· | ···················· | ···················· | ···················· |
| P | 3173 hCoV-19/Denmark/DCGC-2851/2020\|EPI_ISL_616678\|2020 | ···················· | ···················· | ···················· | ···················· | ···················· | ···················· |
| P | 3174 hCoV-19/Denmark/DCGC-2929/2020\|EPI_ISL_616679\|2020 | ···················· | ·A·················· | ···················· | ···················· | ···················· | ··A················· |
| P | 3175 hCoV-19/Denmark/DCGC-2852/2020\|EPI_ISL_616680\|2020 | ···················· | ·A·················· | ···················· | ···················· | ···················· | ··A················· |
| P | 3176 hCoV-19/Denmark/DCGC-2853/2020\|EPI_ISL_616681\|2020 | ···················· | ···················· | ···················· | ···················· | ···················· | ···················· |
| P | 3177 hCoV-19/Denmark/DCGC-2854/2020\|EPI_ISL_616682\|2020 | ···················· | ···················· | ···················· | ···················· | ···················· | ···················· |
| P | 3178 hCoV-19/Denmark/DCGC-2855/2020\|EPI_ISL_616683\|2020 | ···················· | ···················· | ···················· | ···················· | ···················· | ···················· |
| P | 3179 hCoV-19/Denmark/DCGC-2856/2020\|EPI_ISL_616684\|2020 | ···················· | ···················· | ···················· | ···················· | ···················· | ···················· |
| P | 3180 hCoV-19/Denmark/DCGC-2857/2020\|EPI_ISL_616685\|2020 | ···················· | ···················· | ···················· | ···················· | ···················· | ···················· |
| P | 3181 hCoV-19/Denmark/DCGC-2858/2020\|EPI_ISL_616686\|2020 | ···················· | ···················· | ···················· | ···················· | ···················· | ···················· |
| P | 3182 hCoV-19/Denmark/DCGC-2859/2020\|EPI_ISL_616687\|2020 | ···················· | ···················· | ···················· | ···················· | ···················· | ···················· |
| P | 3183 hCoV-19/Denmark/DCGC-2861/2020\|EPI_ISL_616689\|2020 | ···················· | ···················· | ···················· | ···················· | ···················· | ···················· |
| P | 3184 hCoV-19/Denmark/DCGC-2862/2020\|EPI_ISL_616690\|2020 | ···················· | ···················· | ···················· | ···················· | ···················· | ···················· |
| P | 3185 hCoV-19/Denmark/DCGC-2863/2020\|EPI_ISL_616691\|2020 | ···················· | ···················· | ···················· | ···················· | ·····T·············· | ···················· |
| P | 3186 hCoV-19/Denmark/DCGC-2864/2020\|EPI_ISL_616692\|2020 | ···················· | ···················· | ···················· | ···················· | ···················· | ···················· |
| P | 3187 hCoV-19/Denmark/DCGC-2933/2020\|EPI_ISL_616694\|2020 | ···················· | ···················· | ···················· | ···················· | ···················· | ···················· |
| P | 3188 hCoV-19/Denmark/DCGC-2935/2020\|EPI_ISL_616696\|2020 | ···················· | ···················· | ···················· | ···················· | ···················· | ···················· |
| P | 3189 hCoV-19/Denmark/DCGC-2937/2020\|EPI_ISL_616698\|2020 | ···················· | ···················· | ···················· | ···················· | ···················· | ···················· |
| P | 3190 hCoV-19/Denmark/DCGC-2938/2020\|EPI_ISL_616699\|2020 | ···················· | ···················· | ···················· | ···················· | ···················· | ···················· |
| P | 3191 hCoV-19/Denmark/DCGC-8452/2020\|EPI_ISL_617927\|2020 | ···················· | ···················· | ···················· | ···················· | ···················· | ···················· |
| P | 3192 hCoV-19/Denmark/DCGC-8460/2020\|EPI_ISL_617928\|2020 | ···················· | ···················· | ···················· | ···················· | ···················· | ···················· |
| P | 3193 hCoV-19/Denmark/DCGC-8465/2020\|EPI_ISL_617929\|2020 | ···················· | ···················· | ···················· | ···················· | ···················· | ···················· |
| P | 3194 hCoV-19/Denmark/DCGC-8472/2020\|EPI_ISL_617930\|2020 | ···················· | ·A·················· | ···················· | ···················· | ···················· | ···················· |
| P | 3195 hCoV-19/Denmark/DCGC-8415/2020\|EPI_ISL_617931\|2020 | ···················· | ···················· | ···················· | ···················· | ···················· | ···················· |
| P | 3196 hCoV-19/Denmark/DCGC-8427/2020\|EPI_ISL_617933\|2020 | ···················· | ···················· | ···················· | ···················· | ···················· | ···················· |
| P | 3197 hCoV-19/Denmark/DCGC-8440/2020\|EPI_ISL_617935\|2020 | ···················· | ···················· | ···················· | ···················· | ···················· | ···················· |
| P | 3198 hCoV-19/Denmark/DCGC-8446/2020\|EPI_ISL_617936\|2020 | ···················· | ···················· | ···················· | ···················· | ···················· | ···················· |
| P | 3199 hCoV-19/Denmark/DCGC-8453/2020\|EPI_ISL_617937\|2020 | ···················· | ·A·················· | ···················· | ···················· | ···················· | ···················· |
| P | 3200 hCoV-19/Denmark/DCGC-8461/2020\|EPI_ISL_617938\|2020 | ···················· | ···················· | ···················· | ···················· | ···················· | ···················· |
| P | 3201 hCoV-19/Denmark/DCGC-1816/2020\|EPI_ISL_617939\|2020 | ···················· | ···················· | ···················· | ···················· | ···················· | ···················· |
| P | 3202 hCoV-19/Denmark/DCGC-1345/2020\|EPI_ISL_617944\|2020 | ···················· | ···················· | ···················· | ···················· | ···················· | ···················· |
| P | 3203 hCoV-19/Denmark/DCGC-1726/2020\|EPI_ISL_617945\|2020 | ···················· | ···················· | ···················· | ···················· | ···················· | ···················· |
| P | 3204 hCoV-19/Denmark/DCGC-1727/2020\|EPI_ISL_617946\|2020 | ···················· | ···················· | ···················· | ···················· | ···················· | ···················· |
| P | 3205 hCoV-19/Denmark/DCGC-1728/2020\|EPI_ISL_617947\|2020 | ···················· | ···················· | ···················· | ···················· | ···················· | ···················· |
| P | 3206 hCoV-19/Denmark/DCGC-1729/2020\|EPI_ISL_617948\|2020 | ···················· | ···················· | ···················· | ···················· | ···················· | ···················· |
| P | 3207 hCoV-19/Denmark/DCGC-1538/2020\|EPI_ISL_617990\|2020 | ···················· | ···················· | ···················· | ···················· | ···················· | ···················· |
| P | 3208 hCoV-19/Denmark/DCGC-1295/2020\|EPI_ISL_617991\|2020 | ···················· | ···················· | ···················· | ···················· | ···················· | ···················· |
| P | 3209 hCoV-19/Denmark/DCGC-1311/2020\|EPI_ISL_617992\|2020 | ···················· | ···················· | ···················· | ···················· | ···················· | ···················· |
| P | 3210 hCoV-19/Denmark/DCGC-1708/2020\|EPI_ISL_617993\|2020 | ···················· | ···················· | ···················· | ···················· | ···················· | ···················· |
| P | 3211 hCoV-19/Denmark/DCGC-1321/2020\|EPI_ISL_617994\|2020 | ···················· | ···················· | ···················· | ···················· | ···················· | ···················· |
| P | 3212 hCoV-19/Denmark/DCGC-1322/2020\|EPI_ISL_617995\|2020 | ···················· | ···················· | ···················· | ···················· | ···················· | ···················· |
| P | 3213 hCoV-19/Denmark/DCGC-1323/2020\|EPI_ISL_617996\|2020 | ···················· | ···················· | ···················· | ···················· | ···················· | ···················· |
| P | 3214 hCoV-19/Denmark/DCGC-1324/2020\|EPI_ISL_617997\|2020 | ···················· | ···················· | ···················· | ···················· | ···················· | ···················· |
| P | 3215 hCoV-19/Denmark/DCGC-1325/2020\|EPI_ISL_617998\|2020 | ···················· | ···················· | ···················· | ···················· | ···················· | ···················· |
| P | 3216 hCoV-19/Denmark/DCGC-1326/2020\|EPI_ISL_617999\|2020 | ···················· | ···················· | ···················· | ···················· | ···················· | ···················· |
| P | 3217 hCoV-19/Denmark/DCGC-6141/2020\|EPI_ISL_621175\|2020 | ···················· | ···················· | ···················· | ···················· | ···················· | ···················· |
| P | 3218 hCoV-19/Denmark/DCGC-6142/2020\|EPI_ISL_621176\|2020 | ···················· | ···················· | ···················· | ···················· | ···················· | ···················· |
| P | 3219 hCoV-19/Denmark/DCGC-6334/2020\|EPI_ISL_621191\|2020 | ···················· | ·A·················· | ···················· | ···················· | ···················· | ··A················· |
| P | 3220 hCoV-19/Denmark/DCGC-6335/2020\|EPI_ISL_621192\|2020 | ···················· | ···················· | ···················· | ···················· | ···················· | ···················· |
| P | 3221 hCoV-19/Denmark/DCGC-6337/2020\|EPI_ISL_621194\|2020 | ···················· | ···················· | ···················· | ···················· | ···················· | ···················· |
| P | 3222 hCoV-19/Denmark/DCGC-5690/2020\|EPI_ISL_621195\|2020 | ···················· | ···················· | ···················· | ···················· | ···················· | ···················· |
| P | 3223 hCoV-19/Denmark/DCGC-5691/2020\|EPI_ISL_621196\|2020 | ···················· | ···················· | ···················· | ···················· | ···················· | ···················· |
| P | 3224 hCoV-19/Denmark/DCGC-5692/2020\|EPI_ISL_621197\|2020 | ···················· | ···················· | ···················· | ···················· | ···················· | ···················· |
| P | 3225 hCoV-19/Denmark/DCGC-5693/2020\|EPI_ISL_621198\|2020 | ···················· | ···················· | ···················· | ···················· | ···················· | ···················· |
| P | 3226 hCoV-19/Denmark/DCGC-5694/2020\|EPI_ISL_621199\|2020 | ···················· | ···················· | ···················· | ···················· | ···················· | ···················· |
| P | 3227 hCoV-19/Denmark/DCGC-6748/2020\|EPI_ISL_622400\|2020 | ···················· | ···················· | ···················· | ···················· | ···················· | ···················· |
| P | 3228 hCoV-19/Denmark/DCGC-6753/2020\|EPI_ISL_622401\|2020 | ···················· | ···················· | ···················· | ···················· | ···················· | ···················· |
| P | 3229 hCoV-19/Denmark/DCGC-6756/2020\|EPI_ISL_622402\|2020 | ···················· | ···················· | ···················· | ···················· | ···················· | ···················· |
| P | 3230 hCoV-19/Denmark/DCGC-1654/2020\|EPI_ISL_622403\|2020 | ···················· | ···················· | ···················· | ···················· | ···················· | ···················· |
| P | 3231 hCoV-19/Denmark/DCGC-1658/2020\|EPI_ISL_622407\|2020 | ···················· | ···················· | ···················· | ···················· | ···················· | ···················· |
| P | 3232 hCoV-19/Denmark/DCGC-1659/2020\|EPI_ISL_622408\|2020 | ···················· | ···················· | ···················· | ···················· | ···················· | ···················· |
| P | 3233 hCoV-19/Denmark/DCGC-1661/2020\|EPI_ISL_622410\|2020 | ···················· | ···················· | ···················· | ···················· | ···················· | ···················· |
| P | 3234 hCoV-19/Denmark/DCGC-1662/2020\|EPI_ISL_622411\|2020 | ···················· | ···················· | ···················· | ···················· | ···················· | ···················· |
| P | 3235 hCoV-19/Denmark/DCGC-1663/2020\|EPI_ISL_622412\|2020 | ···················· | ···················· | ···················· | ···················· | ···················· | ···················· |
| P | 3236 hCoV-19/Denmark/DCGC-1664/2020\|EPI_ISL_622413\|2020 | ···················· | ···················· | ···················· | ···················· | ···················· | ···················· |
| P | 3237 hCoV-19/Denmark/DCGC-1665/2020\|EPI_ISL_622414\|2020 | ···················· | ···················· | ···················· | ···················· | ···················· | ···················· |

```
P 3238 hCoV-19/England/CAMC-AB23FB/2020|EPI_ISL_623702|20 ···················· ···················· ···················· ···················· ···················· ····················
P 3239 hCoV-19/England/CAMC-AB222B/2020|EPI_ISL_623703|20 ···················· ···················· ···················· ···················· ···················· ····················
P 3240 hCoV-19/England/MILK-AB8D42/2020|EPI_ISL_623704|20 ···················· ···················· ···················· ···················· ···················· ····················
P 3241 hCoV-19/England/MILK-AB8CFA/2020|EPI_ISL_623705|20 ···················· ···················· ···················· ···················· ···················· ····················
P 3242 hCoV-19/England/MILK-AB9BED/2020|EPI_ISL_623706|20 ···················· ···················· ···················· ···················· ···················· ····················
P 3243 hCoV-19/England/MILK-AB8762/2020|EPI_ISL_623707|20 ···················· ···················· ···················· ···················· ···················· ····················
P 3244 hCoV-19/England/MILK-AB93A9/2020|EPI_ISL_623708|20 ···················· ···················· ···················· ···················· ···················· ····················
P 3245 hCoV-19/England/MILK-AB8E6D/2020|EPI_ISL_623709|20 ···················· ···················· ···················· ···················· ······T············· ····················
P 3246 hCoV-19/England/MILK-AB87BD/2020|EPI_ISL_623711|20 ···················· ···················· ···················· ···················· ···················· ····················
P 3247 hCoV-19/England/CAMC-AB1C00/2020|EPI_ISL_623712|20 ···················· ···················· ···················· ···················· ······T············· ····················
P 3248 hCoV-19/England/MILK-AB8E7C/2020|EPI_ISL_623714|20 ···················· ···················· ···················· ···················· ···················· ····················
P 3249 hCoV-19/England/MILK-AB7BA3/2020|EPI_ISL_623715|20 ···················· ···················· ···················· ···················· ···················· ····················
P 3250 hCoV-19/England/CAMC-AB13ED/2020|EPI_ISL_623717|20 ···················· ···················· ···················· ···················· ···················· ····················
P 3251 hCoV-19/England/MILK-AB8735/2020|EPI_ISL_623718|20 ···················· ···················· ···················· ···················· ···················· ····················
P 3252 hCoV-19/England/MILK-AB7B2B/2020|EPI_ISL_623720|20 ···················· ···················· ···················· ···················· ···················· ····················
P 3253 hCoV-19/England/CAMC-AB15C9/2020|EPI_ISL_623721|20 ···················· ···················· ···················· ···················· ···················· ····················
P 3254 hCoV-19/England/MILK-AB7E7D/2020|EPI_ISL_623722|20 ···················· ·A·················· ···················· ···················· ···················· ···A················
P 3255 hCoV-19/England/MILK-AB8D8E/2020|EPI_ISL_623723|20 ···················· ···················· ···················· ···················· ···················· ····················
P 3256 hCoV-19/England/CAMC-AB1E0A/2020|EPI_ISL_623725|20 ···················· ···················· ···················· ···················· ···················· ····················
P 3257 hCoV-19/England/CAMC-AB18FD/2020|EPI_ISL_623726|20 ···················· ···················· ···················· ···················· ···················· ····················
P 3258 hCoV-19/England/MILK-AB7ED7/2020|EPI_ISL_623727|20 ···················· ·A·················· ···················· ···················· ···················· ····················
P 3259 hCoV-19/England/MILK-AB902A/2020|EPI_ISL_623728|20 ···················· ·A·················· ···················· ···················· ···················· ···A················
P 3260 hCoV-19/England/MILK-AB78AC/2020|EPI_ISL_623729|20 ···················· ···················· ···················· ···················· ···················· ····················
P 3261 hCoV-19/England/CAMC-AB115C/2020|EPI_ISL_623731|20 ···················· ···················· ···················· ···················· ···················· ····················
P 3262 hCoV-19/England/CAMC-AB1EBF/2020|EPI_ISL_623733|20 ···················· ···················· ···················· ···················· ···················· ····················
P 3263 hCoV-19/England/MILK-AB797C/2020|EPI_ISL_623734|20 ···················· ···················· ···················· ···················· ···················· ····················
P 3264 hCoV-19/England/MILK-AB7C91/2020|EPI_ISL_623737|20 ···················· ···················· ···················· ···················· ···················· ····················
P 3265 hCoV-19/England/MILK-AB7B1C/2020|EPI_ISL_623739|20 ···················· ···················· ···················· ···················· ···················· ····················
P 3266 hCoV-19/England/CAMC-AB23EC/2020|EPI_ISL_623740|20 ···················· ···················· ···················· ···················· ···················· ····················
P 3267 hCoV-19/England/CAMC-AB14AE/2020|EPI_ISL_623742|20 ···················· ···················· ···················· ···················· ···················· ····················
P 3268 hCoV-19/England/MILK-AB79F4/2020|EPI_ISL_623744|20 ···················· ···················· ···················· ···················· ···················· ····················
P 3269 hCoV-19/England/MILK-AB79E5/2020|EPI_ISL_623745|20 ···················· ···················· ···················· ···················· ···················· ····················
P 3270 hCoV-19/Denmark/DCGC-8625/2020|EPI_ISL_626000|2020 ···················· ···················· ···················· ···················· ······T············· ····················
P 3271 hCoV-19/Denmark/DCGC-8628/2020|EPI_ISL_626001|2020 ···················· ···················· ···················· ···················· ···················· ····················
P 3272 hCoV-19/Denmark/DCGC-8635/2020|EPI_ISL_626002|2020 ···················· ···················· ···················· ···················· ···················· ····················
P 3273 hCoV-19/Denmark/DCGC-8640/2020|EPI_ISL_626003|2020 ···················· ···················· ···················· ···················· ···················· ····················
P 3274 hCoV-19/Denmark/DCGC-8664/2020|EPI_ISL_626004|2020 ···················· ·A·················· ···················· ···················· ···················· ····················
P 3275 hCoV-19/Denmark/DCGC-8672/2020|EPI_ISL_626005|2020 ···················· ···················· ···················· ···················· ···················· ····················
P 3276 hCoV-19/Denmark/DCGC-8636/2020|EPI_ISL_626006|2020 ···················· ···················· ···················· ···················· ···················· ····················
P 3277 hCoV-19/Denmark/DCGC-8641/2020|EPI_ISL_626007|2020 ···················· ···················· ···················· ···················· ···················· ····················
P 3278 hCoV-19/Denmark/DCGC-8660/2020|EPI_ISL_626008|2020 ···················· ·A·················· ···················· ···················· ···················· ····················
P 3279 hCoV-19/Denmark/DCGC-8665/2020|EPI_ISL_626009|2020 ···················· ·A·················· ···················· ···················· ···················· ····················
P 3280 hCoV-19/Denmark/DCGC-8695/2020|EPI_ISL_626011|2020 ···················· ···················· ···················· ···················· ······T············· ····················
P 3281 hCoV-19/Denmark/DCGC-8679/2020|EPI_ISL_626012|2020 ···················· ···················· ···················· ···················· ······T············· ····················
P 3282 hCoV-19/Denmark/DCGC-8696/2020|EPI_ISL_626013|2020 ···················· ···················· ···················· ···················· ···················· ····················
P 3283 hCoV-19/Denmark/DCGC-8701/2020|EPI_ISL_626014|2020 ···················· ···················· ···················· ···················· ···················· ····················
P 3284 hCoV-19/Denmark/DCGC-8719/2020|EPI_ISL_626015|2020 ···················· ···················· ···················· ···················· ···················· ····················
P 3285 hCoV-19/Denmark/DCGC-8730/2020|EPI_ISL_626016|2020 ···················· ···················· ···················· ···················· ···················· ····················
P 3286 hCoV-19/Denmark/DCGC-8731/2020|EPI_ISL_626017|2020 ···················· ···················· ···················· ···················· ···················· ····················
P 3287 hCoV-19/Denmark/DCGC-8680/2020|EPI_ISL_626018|2020 ···················· ···················· ···················· ···················· ···················· ····················
P 3288 hCoV-19/Denmark/DCGC-8720/2020|EPI_ISL_626019|2020 ···················· ···················· ···················· ···················· ···················· ····················
P 3289 hCoV-19/Denmark/DCGC-8741/2020|EPI_ISL_626020|2020 ···················· ···················· ···················· ···················· ···················· ····················
P 3290 hCoV-19/Denmark/DCGC-8686/2020|EPI_ISL_626021|2020 ···················· ·A·················· ···················· ···················· ···················· ····················
P 3291 hCoV-19/Denmark/DCGC-8691/2020|EPI_ISL_626022|2020 ···················· ···················· ···················· ···················· ···················· ····················
P 3292 hCoV-19/Denmark/DCGC-8697/2020|EPI_ISL_626023|2020 ···················· ···················· ···················· ···················· ···················· ····················
P 3293 hCoV-19/Denmark/DCGC-8710/2020|EPI_ISL_626024|2020 ···················· ···················· ···················· ···················· ···················· ····················
P 3294 hCoV-19/Denmark/DCGC-8726/2020|EPI_ISL_626025|2020 ···················· ···················· ···················· ···················· ···················· ····················
P 3295 hCoV-19/Denmark/DCGC-8732/2020|EPI_ISL_626026|2020 ···················· ···················· ···················· ···················· ···················· ····················
P 3296 hCoV-19/Denmark/DCGC-8711/2020|EPI_ISL_626027|2020 ···················· ···················· ···················· ···················· ···················· ····················
P 3297 hCoV-19/Denmark/DCGC-8716/2020|EPI_ISL_626028|2020 ········T··········· ···················· ···················· ···················· ···················· ····················
P 3298 hCoV-19/Denmark/DCGC-8683/2020|EPI_ISL_626029|2020 ···················· ·A·················· ···················· ···················· ···················· ···A················
P 3299 hCoV-19/Denmark/DCGC-8693/2020|EPI_ISL_626030|2020 ···················· ···················· ···················· ···················· ···················· ····················
P 3300 hCoV-19/Denmark/DCGC-8698/2020|EPI_ISL_626031|2020 ···················· ···················· ···················· ···················· ···················· ····················
P 3301 hCoV-19/Denmark/DCGC-8717/2020|EPI_ISL_626032|2020 ···················· ···················· ···················· ···················· ···················· ····················
P 3302 hCoV-19/Denmark/DCGC-8706/2020|EPI_ISL_626033|2020 ···················· ···················· ···················· ···················· ···················· ····················
P 3303 hCoV-19/Denmark/DCGC-8734/2020|EPI_ISL_626034|2020 ···················· ···················· ···················· ···················· ···················· ····················
P 3304 hCoV-19/Denmark/DCGC-8742/2020|EPI_ISL_626035|2020 ···················· ···················· ···················· ···················· ···················· ····················
P 3305 hCoV-19/Denmark/DCGC-8751/2020|EPI_ISL_626036|2020 ···················· ···················· ···················· ···················· ···················· ····················
P 3306 hCoV-19/Denmark/DCGC-8757/2020|EPI_ISL_626037|2020 ···················· ···················· ···················· ···················· ···················· ····················
P 3307 hCoV-19/Denmark/DCGC-8768/2020|EPI_ISL_626038|2020 ···················· ···················· ···················· ···················· ···················· ····················
P 3308 hCoV-19/Denmark/DCGC-8775/2020|EPI_ISL_626039|2020 ···················· ···················· ···················· ···················· ···················· ····················
P 3309 hCoV-19/Denmark/DCGC-8793/2020|EPI_ISL_626040|2020 ···················· ···················· ···················· ···················· ···················· ····················
P 3310 hCoV-19/Denmark/DCGC-8801/2020|EPI_ISL_626041|2020 ···················· ···················· ···················· ···················· ···················· ····················
P 3311 hCoV-19/Denmark/DCGC-8743/2020|EPI_ISL_626042|2020 ···················· ···················· ···················· ···················· ···················· ····················
P 3312 hCoV-19/Denmark/DCGC-8769/2020|EPI_ISL_626043|2020 ···················· ···················· ···················· ···················· ···················· ····················
P 3313 hCoV-19/Denmark/DCGC-8776/2020|EPI_ISL_626044|2020 ···················· ···················· ···················· ···················· ···················· ····················
P 3314 hCoV-19/Denmark/DCGC-8781/2020|EPI_ISL_626045|2020 ···················· ···················· ···················· ···················· ···················· ····················
P 3315 hCoV-19/Denmark/DCGC-8783/2020|EPI_ISL_626046|2020 ···················· ···················· ···················· ···················· ···················· ····················
P 3316 hCoV-19/Denmark/DCGC-8794/2020|EPI_ISL_626047|2020 ···················· ·A·················· ···················· ···················· ···················· ···A················
P 3317 hCoV-19/Denmark/DCGC-8770/2020|EPI_ISL_626048|2020 ···················· ···················· ···················· ···················· ···················· ····················
```

| | | | | | | | | |
|---|---|---|---|---|---|---|---|---|
| P | 3318 | hCoV-19/Denmark/DCGC-8782/2020\|EPI_ISL_626049\|2020 | ···················· | ···················· | ···················· | ···················· | ···················· | ···················· |
| P | 3319 | hCoV-19/Denmark/DCGC-8803/2020\|EPI_ISL_626050\|2020 | ···················· | ·A·················· | ···················· | ···················· | ···················· | ···················· |
| P | 3320 | hCoV-19/Denmark/DCGC-8753/2020\|EPI_ISL_626051\|2020 | ···················· | ···················· | ···················· | ···················· | ···················· | ···················· |
| P | 3321 | hCoV-19/Denmark/DCGC-8759/2020\|EPI_ISL_626052\|2020 | ···················· | ···················· | ···················· | ···················· | ···················· | ···················· |
| P | 3322 | hCoV-19/Denmark/DCGC-8778/2020\|EPI_ISL_626053\|2020 | ···················· | ···················· | ···················· | ···················· | ···················· | ···················· |
| P | 3323 | hCoV-19/Denmark/DCGC-8784/2020\|EPI_ISL_626054\|2020 | ···················· | ···················· | ···················· | ···················· | ···················· | ···················· |
| P | 3324 | hCoV-19/Denmark/DCGC-8796/2020\|EPI_ISL_626055\|2020 | ···················· | ···················· | ···················· | ···················· | ···················· | ···················· |
| P | 3325 | hCoV-19/Denmark/DCGC-8760/2020\|EPI_ISL_626056\|2020 | ···················· | ···················· | ···················· | ···················· | ···················· | ···················· |
| P | 3326 | hCoV-19/Denmark/DCGC-8779/2020\|EPI_ISL_626057\|2020 | ···················· | ···················· | ···················· | ···················· | ···················· | ···················· |
| P | 3327 | hCoV-19/Denmark/DCGC-8785/2020\|EPI_ISL_626058\|2020 | ···················· | ···················· | ···················· | ···················· | ···················· | ···················· |
| P | 3328 | hCoV-19/Denmark/DCGC-8804/2020\|EPI_ISL_626059\|2020 | ···················· | ···················· | ···················· | ···················· | ···················· | ···················· |
| P | 3329 | hCoV-19/Denmark/DCGC-5359/2020\|EPI_ISL_615290\|2020 | ···················· | ···················· | ···················· | ···················· | ···················· | ···················· |
| P | 3330 | hCoV-19/Denmark/DCGC-5360/2020\|EPI_ISL_615291\|2020 | ···················· | ···················· | ···················· | ···················· | ···················· | ···················· |
| P | 3331 | hCoV-19/Denmark/DCGC-5361/2020\|EPI_ISL_615292\|2020 | ···················· | ···················· | ···················· | ···················· | ···················· | ···················· |
| P | 3332 | hCoV-19/Denmark/DCGC-5362/2020\|EPI_ISL_615293\|2020 | ···················· | ···················· | ···················· | ···················· | ···················· | ···················· |
| P | 3333 | hCoV-19/Denmark/DCGC-5363/2020\|EPI_ISL_615294\|2020 | ···················· | ···················· | ···················· | ···················· | ···················· | ···················· |
| P | 3334 | hCoV-19/Denmark/DCGC-5364/2020\|EPI_ISL_615295\|2020 | ···················· | ···················· | ···················· | ···················· | ···················· | ···················· |
| P | 3335 | hCoV-19/Denmark/DCGC-5366/2020\|EPI_ISL_615297\|2020 | ···················· | ···················· | ···················· | ···················· | ···················· | ···················· |
| P | 3336 | hCoV-19/Denmark/DCGC-5368/2020\|EPI_ISL_615299\|2020 | ···················· | ···················· | ···················· | ···················· | ···················· | ···················· |
| P | 3337 | hCoV-19/Denmark/DCGC-2621/2020\|EPI_ISL_616500\|2020 | ···················· | ···················· | ···················· | ···················· | ···················· | ···················· |
| P | 3338 | hCoV-19/Denmark/DCGC-2623/2020\|EPI_ISL_616502\|2020 | ···················· | ···················· | ···················· | ···················· | ···················· | ···················· |
| P | 3339 | hCoV-19/Denmark/DCGC-2624/2020\|EPI_ISL_616503\|2020 | ···················· | ···················· | ···················· | ···················· | ···················· | ···················· |
| P | 3340 | hCoV-19/Denmark/DCGC-2625/2020\|EPI_ISL_616504\|2020 | ···················· | ···················· | ···················· | ···················· | ···················· | ···················· |
| P | 3341 | hCoV-19/Denmark/DCGC-2626/2020\|EPI_ISL_616505\|2020 | ···················· | ···················· | ···················· | ···················· | ···················· | ···················· |
| P | 3342 | hCoV-19/Denmark/DCGC-2627/2020\|EPI_ISL_616506\|2020 | ···················· | ···················· | ···················· | ···················· | ···················· | ···················· |
| P | 3343 | hCoV-19/Denmark/DCGC-2628/2020\|EPI_ISL_616507\|2020 | ···················· | ···················· | ···················· | ···················· | ···················· | ···················· |
| P | 3344 | hCoV-19/Denmark/DCGC-2629/2020\|EPI_ISL_616508\|2020 | ···················· | ···················· | ···················· | ···················· | ···················· | ···················· |
| P | 3345 | hCoV-19/Denmark/DCGC-2630/2020\|EPI_ISL_616509\|2020 | ···················· | ···················· | ···················· | ···················· | ···················· | ···················· |
| P | 3346 | hCoV-19/Denmark/DCGC-2631/2020\|EPI_ISL_616510\|2020 | ···················· | ···················· | ···················· | ···················· | ···················· | ···················· |
| P | 3347 | hCoV-19/Denmark/DCGC-2632/2020\|EPI_ISL_616511\|2020 | ···················· | ·A·················· | ···················· | ···················· | ···················· | ··A················· |
| P | 3348 | hCoV-19/Denmark/DCGC-2633/2020\|EPI_ISL_616512\|2020 | ···················· | ·A·················· | ···················· | ···················· | ···················· | ··A················· |
| P | 3349 | hCoV-19/Denmark/DCGC-2634/2020\|EPI_ISL_616513\|2020 | ···················· | ···················· | ···················· | ···················· | ···················· | ···················· |
| P | 3350 | hCoV-19/Denmark/DCGC-2635/2020\|EPI_ISL_616514\|2020 | ···················· | ···················· | ···················· | ···················· | ···················· | ···················· |
| P | 3351 | hCoV-19/Denmark/DCGC-2636/2020\|EPI_ISL_616515\|2020 | ···················· | ···················· | ···················· | ···················· | ···················· | ···················· |
| P | 3352 | hCoV-19/Denmark/DCGC-2637/2020\|EPI_ISL_616516\|2020 | ···················· | ···················· | ···················· | ···················· | ···················· | ···················· |
| P | 3353 | hCoV-19/Denmark/DCGC-2638/2020\|EPI_ISL_616517\|2020 | ···················· | ···················· | ···················· | ···················· | ···················· | ···················· |
| P | 3354 | hCoV-19/Denmark/DCGC-2542/2020\|EPI_ISL_616518\|2020 | ···················· | ·A·················· | ···················· | ···················· | ···················· | ···················· |
| P | 3355 | hCoV-19/Denmark/DCGC-2546/2020\|EPI_ISL_616522\|2020 | ···················· | ···················· | ···················· | ···················· | ···················· | ···················· |
| P | 3356 | hCoV-19/Denmark/DCGC-2547/2020\|EPI_ISL_616523\|2020 | ···················· | ···················· | ···················· | ···················· | ···················· | ···················· |
| P | 3357 | hCoV-19/Denmark/DCGC-2548/2020\|EPI_ISL_616524\|2020 | ···················· | ·A··A··············· | ···················· | ···················· | ···················· | ···················· |
| P | 3358 | hCoV-19/Denmark/DCGC-2549/2020\|EPI_ISL_616525\|2020 | ···················· | ·A··A··············· | ···················· | ···················· | ···················· | ···················· |
| P | 3359 | hCoV-19/Denmark/DCGC-2553/2020\|EPI_ISL_616529\|2020 | ·······T············ | ···················· | ···················· | ···················· | ···················· | ···················· |
| P | 3360 | hCoV-19/Denmark/DCGC-2592/2020\|EPI_ISL_617828\|2020 | ···················· | ···················· | ···················· | ···················· | ···················· | ···················· |
| P | 3361 | hCoV-19/Denmark/DCGC-2593/2020\|EPI_ISL_617829\|2020 | ···················· | ···················· | ···················· | ···················· | ···················· | ···················· |
| P | 3362 | hCoV-19/Denmark/DCGC-2594/2020\|EPI_ISL_617830\|2020 | ···················· | ·A·················· | ···················· | ···················· | ···················· | ···················· |
| P | 3363 | hCoV-19/Denmark/DCGC-2595/2020\|EPI_ISL_617831\|2020 | ···················· | ···················· | ···················· | ···················· | ···················· | ···················· |
| P | 3364 | hCoV-19/Denmark/DCGC-2596/2020\|EPI_ISL_617832\|2020 | ···················· | ···················· | ···················· | ···················· | ···················· | ···················· |
| P | 3365 | hCoV-19/Denmark/DCGC-2597/2020\|EPI_ISL_617833\|2020 | ···················· | ···················· | ···················· | ···················· | ···················· | ···················· |
| P | 3366 | hCoV-19/Denmark/DCGC-2598/2020\|EPI_ISL_617834\|2020 | ···················· | ···················· | ···················· | ···················· | ···················· | ···················· |
| P | 3367 | hCoV-19/Denmark/DCGC-2599/2020\|EPI_ISL_617835\|2020 | ···················· | ···················· | ···················· | ···················· | ···················· | ···················· |
| P | 3368 | hCoV-19/Denmark/DCGC-2600/2020\|EPI_ISL_617836\|2020 | ···················· | ···················· | ···················· | ···················· | ···················· | ···················· |
| P | 3369 | hCoV-19/Denmark/DCGC-2601/2020\|EPI_ISL_617837\|2020 | ···················· | ···················· | ···················· | ···················· | ···················· | ···················· |
| P | 3370 | hCoV-19/Denmark/DCGC-2602/2020\|EPI_ISL_617838\|2020 | ···················· | ···················· | ···················· | ···················· | ···················· | ···················· |
| P | 3371 | hCoV-19/Denmark/DCGC-2569/2020\|EPI_ISL_617839\|2020 | ···················· | ···················· | ···················· | ···················· | ···················· | ···················· |
| P | 3372 | hCoV-19/Denmark/DCGC-2603/2020\|EPI_ISL_617840\|2020 | ···················· | ···················· | ···················· | ···················· | ···················· | ···················· |
| P | 3373 | hCoV-19/Denmark/DCGC-2604/2020\|EPI_ISL_617841\|2020 | ···················· | ···················· | ···················· | ···················· | ···················· | ···················· |
| P | 3374 | hCoV-19/Denmark/DCGC-2605/2020\|EPI_ISL_617842\|2020 | ···················· | ···················· | ···················· | ···················· | ···················· | ···················· |
| P | 3375 | hCoV-19/Denmark/DCGC-2606/2020\|EPI_ISL_617843\|2020 | ···················· | ···················· | ···················· | ···················· | ···················· | ···················· |
| P | 3376 | hCoV-19/Denmark/DCGC-2607/2020\|EPI_ISL_617844\|2020 | ···················· | ···················· | ···················· | ···················· | ···················· | ···················· |
| P | 3377 | hCoV-19/Denmark/DCGC-2608/2020\|EPI_ISL_617845\|2020 | ···················· | ·A·················· | ···················· | ···················· | ···················· | ···················· |
| P | 3378 | hCoV-19/Denmark/DCGC-2609/2020\|EPI_ISL_617846\|2020 | ···················· | ···················· | ···················· | ···················· | ···················· | ···················· |
| P | 3379 | hCoV-19/Denmark/DCGC-2610/2020\|EPI_ISL_617847\|2020 | ···················· | ···················· | ···················· | ···················· | ···················· | ···················· |
| P | 3380 | hCoV-19/Denmark/DCGC-2611/2020\|EPI_ISL_617848\|2020 | ···················· | ···················· | ···················· | ···················· | ···················· | ···················· |
| P | 3381 | hCoV-19/Denmark/DCGC-2612/2020\|EPI_ISL_617849\|2020 | ···················· | ···················· | ···················· | ···················· | ···················· | ···················· |
| P | 3382 | hCoV-19/Denmark/DCGC-2616/2020\|EPI_ISL_617853\|2020 | ···················· | ···················· | ···················· | ···················· | ···················· | ···················· |
| P | 3383 | hCoV-19/Denmark/DCGC-2620/2020\|EPI_ISL_617857\|2020 | ···················· | ···················· | ···················· | ···················· | ···················· | ···················· |
| P | 3384 | hCoV-19/Denmark/DCGC-2639/2020\|EPI_ISL_617858\|2020 | ···················· | ···················· | ···················· | ···················· | ···················· | ···················· |
| P | 3385 | hCoV-19/Denmark/DCGC-2640/2020\|EPI_ISL_617859\|2020 | ···················· | ···················· | ···················· | ···················· | ···················· | ···················· |
| P | 3386 | hCoV-19/Denmark/DCGC-2679/2020\|EPI_ISL_617900\|2020 | ···················· | ···················· | ···················· | ···················· | ···················· | ···················· |
| P | 3387 | hCoV-19/Denmark/DCGC-2680/2020\|EPI_ISL_617901\|2020 | ···················· | ···················· | ···················· | ···················· | ···················· | ···················· |
| P | 3388 | hCoV-19/Denmark/DCGC-2681/2020\|EPI_ISL_617902\|2020 | ···················· | ···················· | ···················· | ···················· | ···················· | ···················· |
| P | 3389 | hCoV-19/Denmark/DCGC-2682/2020\|EPI_ISL_617903\|2020 | ···················· | ···················· | ···················· | ···················· | ···················· | ···················· |
| P | 3390 | hCoV-19/Denmark/DCGC-2683/2020\|EPI_ISL_617904\|2020 | ···················· | ···················· | ···················· | ···················· | ···················· | ···················· |
| P | 3391 | hCoV-19/Denmark/DCGC-5981/2020\|EPI_ISL_621090\|2020 | ···················· | ···················· | ···················· | ···················· | ···················· | ···················· |
| P | 3392 | hCoV-19/Denmark/DCGC-5982/2020\|EPI_ISL_621091\|2020 | ···················· | ···················· | ···················· | ···················· | ···················· | ···················· |
| P | 3393 | hCoV-19/Denmark/DCGC-5983/2020\|EPI_ISL_621092\|2020 | ···················· | ···················· | ···················· | ···················· | ···················· | ···················· |
| P | 3394 | hCoV-19/Denmark/DCGC-5984/2020\|EPI_ISL_621093\|2020 | ···················· | ···················· | ···················· | ···················· | ···················· | ···················· |
| P | 3395 | hCoV-19/Denmark/DCGC-5985/2020\|EPI_ISL_621094\|2020 | ···················· | ···················· | ···················· | ···················· | ···················· | ···················· |
| P | 3396 | hCoV-19/Denmark/DCGC-5986/2020\|EPI_ISL_621095\|2020 | ···················· | ···················· | ···················· | ···················· | ···················· | ···················· |
| P | 3397 | hCoV-19/Denmark/DCGC-5987/2020\|EPI_ISL_621096\|2020 | ···················· | ···················· | ···················· | ···················· | ···················· | ···················· |

| | | | | | | | | |
|---|---|---|---|---|---|---|---|---|
| P | 3398 | hCoV-19/Denmark/DCGC-5988/2020\|EPI_ISL_621097\|2020 | ···················· | ···················· | ····················· | ···················· | ···················· | ···················· |
| P | 3399 | hCoV-19/Denmark/DCGC-5989/2020\|EPI_ISL_621098\|2020 | ···················· | ···················· | ····················· | ···················· | ···················· | ···················· |
| P | 3400 | hCoV-19/Denmark/DCGC-5990/2020\|EPI_ISL_621099\|2020 | ···················· | ···················· | ····················· | ···················· | ···················· | ···················· |
| P | 3401 | hCoV-19/Denmark/DCGC-5991/2020\|EPI_ISL_621100\|2020 | ···················· | ···················· | ····················· | ···················· | ···················· | ···················· |
| P | 3402 | hCoV-19/Denmark/DCGC-5992/2020\|EPI_ISL_621101\|2020 | ···················· | ···················· | ····················· | ···················· | ···················· | ···················· |
| P | 3403 | hCoV-19/Denmark/DCGC-5993/2020\|EPI_ISL_621102\|2020 | ···················· | ···················· | ····················· | ···················· | ···················· | ···················· |
| P | 3404 | hCoV-19/Denmark/DCGC-5994/2020\|EPI_ISL_621103\|2020 | ···················· | ···················· | ····················· | ···················· | ···················· | ···················· |
| P | 3405 | hCoV-19/Denmark/DCGC-5995/2020\|EPI_ISL_621104\|2020 | ···················· | ···················· | ····················· | ···················· | ···················· | ···················· |
| P | 3406 | hCoV-19/Denmark/DCGC-5996/2020\|EPI_ISL_621105\|2020 | ···················· | ···················· | ····················· | ···················· | ···················· | ···················· |
| P | 3407 | hCoV-19/Denmark/DCGC-5997/2020\|EPI_ISL_621106\|2020 | ···················· | ···················· | ····················· | ···················· | ···················· | ···················· |
| P | 3408 | hCoV-19/Denmark/DCGC-5998/2020\|EPI_ISL_621107\|2020 | ···················· | ···················· | ····················· | ···················· | ···················· | ···················· |
| P | 3409 | hCoV-19/Denmark/DCGC-5999/2020\|EPI_ISL_621108\|2020 | ···················· | ···················· | ····················· | ···················· | ···················· | ···················· |
| P | 3410 | hCoV-19/Denmark/DCGC-6000/2020\|EPI_ISL_621109\|2020 | ···················· | ···················· | ····················· | ···················· | ···················· | ···················· |
| P | 3411 | hCoV-19/Denmark/DCGC-6001/2020\|EPI_ISL_621110\|2020 | ···················· | ···················· | ····················· | ···················· | ···················· | ···················· |
| P | 3412 | hCoV-19/Denmark/DCGC-6002/2020\|EPI_ISL_621111\|2020 | ···················· | ···················· | ····················· | ···················· | ···················· | ···················· |
| P | 3413 | hCoV-19/Denmark/DCGC-6003/2020\|EPI_ISL_621112\|2020 | ···················· | ···················· | ····················· | ···················· | ···················· | ···················· |
| P | 3414 | hCoV-19/Denmark/DCGC-6004/2020\|EPI_ISL_621113\|2020 | ···················· | ···················· | ····················· | ···················· | ···················· | ···················· |
| P | 3415 | hCoV-19/Denmark/DCGC-6005/2020\|EPI_ISL_621114\|2020 | ···················· | ···················· | ····················· | ···················· | ···················· | ···················· |
| P | 3416 | hCoV-19/Denmark/DCGC-6006/2020\|EPI_ISL_621115\|2020 | ···················· | ···················· | ····················· | ···················· | ···················· | ···················· |
| P | 3417 | hCoV-19/Denmark/DCGC-6007/2020\|EPI_ISL_621116\|2020 | ···················· | ···················· | ····················· | ···················· | ···················· | ···················· |
| P | 3418 | hCoV-19/Denmark/DCGC-6008/2020\|EPI_ISL_621117\|2020 | ···················· | ···················· | ····················· | ···················· | ···················· | ···················· |
| P | 3419 | hCoV-19/Denmark/DCGC-6009/2020\|EPI_ISL_621118\|2020 | ···················· | ···················· | ····················· | ···················· | ···················· | ···················· |
| P | 3420 | hCoV-19/Denmark/DCGC-6011/2020\|EPI_ISL_621120\|2020 | ···················· | ···················· | ····················· | ···················· | ···················· | ···················· |
| P | 3421 | hCoV-19/Denmark/DCGC-6012/2020\|EPI_ISL_621121\|2020 | ···················· | ···················· | ····················· | ···················· | ···················· | ···················· |
| P | 3422 | hCoV-19/Denmark/DCGC-6013/2020\|EPI_ISL_621122\|2020 | ···················· | ···················· | ····················· | ···················· | ···················· | ···················· |
| P | 3423 | hCoV-19/Denmark/DCGC-6014/2020\|EPI_ISL_621123\|2020 | ···················· | ···················· | ····················· | ···················· | ···················· | ···················· |
| P | 3424 | hCoV-19/Denmark/DCGC-6015/2020\|EPI_ISL_621124\|2020 | ···················· | ···················· | ····················· | ···················· | ···················· | ···················· |
| P | 3425 | hCoV-19/Denmark/DCGC-6016/2020\|EPI_ISL_621125\|2020 | ···················· | ···················· | ····················· | ···················· | ···················· | ···················· |
| P | 3426 | hCoV-19/Denmark/DCGC-6017/2020\|EPI_ISL_621126\|2020 | ···················· | ···················· | ····················· | ···················· | ···················· | ···················· |
| P | 3427 | hCoV-19/Denmark/DCGC-6020/2020\|EPI_ISL_621129\|2020 | ···················· | ···················· | ····················· | ···················· | ···················· | ···················· |
| P | 3428 | hCoV-19/Denmark/DCGC-6021/2020\|EPI_ISL_621130\|2020 | ···················· | ···················· | ····················· | ···················· | ···················· | ···················· |
| P | 3429 | hCoV-19/Denmark/DCGC-6022/2020\|EPI_ISL_621131\|2020 | ···················· | ···················· | ····················· | ···················· | ···················· | ···················· |
| P | 3430 | hCoV-19/Denmark/DCGC-6024/2020\|EPI_ISL_621133\|2020 | ···················· | ···················· | ····················· | ···················· | ···················· | ···················· |
| P | 3431 | hCoV-19/Denmark/DCGC-6025/2020\|EPI_ISL_621134\|2020 | ···················· | ···················· | ····················· | ···················· | ···················· | ···················· |
| P | 3432 | hCoV-19/Denmark/DCGC-6026/2020\|EPI_ISL_621135\|2020 | ···················· | ···················· | ····················· | ···················· | ···················· | ···················· |
| P | 3433 | hCoV-19/Denmark/DCGC-6027/2020\|EPI_ISL_621136\|2020 | ···················· | ···················· | ····················· | ···················· | ···················· | ···················· |
| P | 3434 | hCoV-19/Denmark/DCGC-6028/2020\|EPI_ISL_621137\|2020 | ···················· | ···················· | ····················· | ···················· | ···················· | ···················· |
| P | 3435 | hCoV-19/Denmark/DCGC-6029/2020\|EPI_ISL_621138\|2020 | ···················· | ···················· | ····················· | ···················· | ···················· | ···················· |
| P | 3436 | hCoV-19/Denmark/DCGC-6030/2020\|EPI_ISL_621139\|2020 | ···················· | ···················· | ····················· | ···················· | ···················· | ···················· |
| P | 3437 | hCoV-19/Denmark/DCGC-6031/2020\|EPI_ISL_621140\|2020 | ···················· | ···················· | ····················· | ···················· | ···················· | ···················· |
| P | 3438 | hCoV-19/Denmark/DCGC-6107/2020\|EPI_ISL_621141\|2020 | ···················· | ···················· | ····················· | ···················· | ···················· | ···················· |
| P | 3439 | hCoV-19/Denmark/DCGC-6108/2020\|EPI_ISL_621142\|2020 | ···················· | ···················· | ····················· | ···················· | ···················· | ···················· |
| P | 3440 | hCoV-19/Denmark/DCGC-6109/2020\|EPI_ISL_621143\|2020 | ···················· | ···················· | ····················· | ···················· | ···················· | ···················· |
| P | 3441 | hCoV-19/Denmark/DCGC-6110/2020\|EPI_ISL_621144\|2020 | ···················· | ···················· | ····················· | ···················· | ···················· | ···················· |
| P | 3442 | hCoV-19/Denmark/DCGC-6111/2020\|EPI_ISL_621145\|2020 | ···················· | ···················· | ····················· | ···················· | ···················· | ···················· |
| P | 3443 | hCoV-19/Denmark/DCGC-6112/2020\|EPI_ISL_621146\|2020 | ···················· | ···················· | ····················· | ···················· | ···················· | ···················· |
| P | 3444 | hCoV-19/Denmark/DCGC-6113/2020\|EPI_ISL_621147\|2020 | ···················· | ···················· | ····················· | ···················· | ···················· | ···················· |
| P | 3445 | hCoV-19/Denmark/DCGC-6116/2020\|EPI_ISL_621150\|2020 | ···················· | ···················· | ····················· | ···················· | ···················· | ···················· |
| P | 3446 | hCoV-19/Denmark/DCGC-6117/2020\|EPI_ISL_621151\|2020 | ···················· | ···················· | ····················· | ···················· | ···················· | ···················· |
| P | 3447 | hCoV-19/Denmark/DCGC-6119/2020\|EPI_ISL_621153\|2020 | ···················· | ···················· | ····················· | ···················· | ···················· | ···················· |
| P | 3448 | hCoV-19/Denmark/DCGC-6120/2020\|EPI_ISL_621154\|2020 | ···················· | ···················· | ····················· | ···················· | ···················· | ···················· |
| P | 3449 | hCoV-19/Denmark/DCGC-6121/2020\|EPI_ISL_621155\|2020 | ···················· | ···················· | ····················· | ···················· | ···················· | ···················· |
| P | 3450 | hCoV-19/Denmark/DCGC-6122/2020\|EPI_ISL_621156\|2020 | ···················· | ···················· | ····················· | ···················· | ···················· | ···················· |
| P | 3451 | hCoV-19/Denmark/DCGC-6123/2020\|EPI_ISL_621157\|2020 | ···················· | ···················· | ····················· | ···················· | ···················· | ···················· |
| P | 3452 | hCoV-19/Denmark/DCGC-6124/2020\|EPI_ISL_621158\|2020 | ···················· | ···················· | ····················· | ···················· | ···················· | ···················· |
| P | 3453 | hCoV-19/Denmark/DCGC-6125/2020\|EPI_ISL_621159\|2020 | ···················· | ···················· | ····················· | ···················· | ···················· | ···················· |
| P | 3454 | hCoV-19/Denmark/DCGC-6126/2020\|EPI_ISL_621160\|2020 | ···················· | ···················· | ····················· | ···················· | ···················· | ···················· |
| P | 3455 | hCoV-19/Denmark/DCGC-6127/2020\|EPI_ISL_621161\|2020 | ···················· | ···················· | ····················· | ···················· | ···················· | ···················· |
| P | 3456 | hCoV-19/Denmark/DCGC-6128/2020\|EPI_ISL_621162\|2020 | ···················· | ···················· | ····················· | ···················· | ···················· | ···················· |
| P | 3457 | hCoV-19/Denmark/DCGC-6129/2020\|EPI_ISL_621163\|2020 | ···················· | ···················· | ····················· | ···················· | ···················· | ···················· |
| P | 3458 | hCoV-19/Denmark/DCGC-6130/2020\|EPI_ISL_621164\|2020 | ···················· | ···················· | ····················· | ···················· | ···················· | ···················· |
| P | 3459 | hCoV-19/Denmark/DCGC-6131/2020\|EPI_ISL_621165\|2020 | ···················· | ···················· | ····················· | ···················· | ···················· | ···················· |
| P | 3460 | hCoV-19/Denmark/DCGC-6132/2020\|EPI_ISL_621166\|2020 | ···················· | ···················· | ····················· | ···················· | ···················· | ···················· |
| P | 3461 | hCoV-19/Denmark/DCGC-6133/2020\|EPI_ISL_621167\|2020 | ···················· | ···················· | ····················· | ···················· | ···················· | ···················· |
| P | 3462 | hCoV-19/Denmark/DCGC-6134/2020\|EPI_ISL_621168\|2020 | ···················· | ···················· | ····················· | ···················· | ···················· | ···················· |
| P | 3463 | hCoV-19/Denmark/DCGC-6135/2020\|EPI_ISL_621169\|2020 | ···················· | ···················· | ····················· | ···················· | ···················· | ···················· |
| P | 3464 | hCoV-19/Denmark/DCGC-1666/2020\|EPI_ISL_622415\|2020 | ········T··········· | ···················· | ···········T········· | ···················· | ···················· | ···················· |
| P | 3465 | hCoV-19/Denmark/DCGC-1667/2020\|EPI_ISL_622416\|2020 | ···················· | ···················· | ····················· | ···················· | ···················· | ···················· |
| P | 3466 | hCoV-19/Denmark/DCGC-1669/2020\|EPI_ISL_622418\|2020 | ···················· | ···················· | ····················· | ···················· | ···················· | ···················· |
| P | 3467 | hCoV-19/Denmark/DCGC-1281/2020\|EPI_ISL_622419\|2020 | ···················· | ···················· | ····················· | ···················· | ···················· | ···················· |
| P | 3468 | hCoV-19/Denmark/DCGC-1389/2020\|EPI_ISL_622420\|2020 | ···················· | ···················· | ····················· | ···················· | ···················· | ···················· |
| P | 3469 | hCoV-19/Denmark/DCGC-1396/2020\|EPI_ISL_622421\|2020 | ···················· | ···················· | ····················· | ···················· | ···················· | ···················· |
| P | 3470 | hCoV-19/Denmark/DCGC-1682/2020\|EPI_ISL_622422\|2020 | ···················· | ···················· | ····················· | ···················· | ···················· | ···················· |
| P | 3471 | hCoV-19/Denmark/DCGC-1404/2020\|EPI_ISL_622423\|2020 | ···················· | ···················· | ····················· | ···················· | ···················· | ···················· |
| P | 3472 | hCoV-19/Denmark/DCGC-1408/2020\|EPI_ISL_622424\|2020 | ···················· | ···················· | ····················· | ···················· | ···················· | ···················· |
| P | 3473 | hCoV-19/Denmark/DCGC-1414/2020\|EPI_ISL_622425\|2020 | ···················· | ···················· | ····················· | ···················· | ···················· | ···················· |
| P | 3474 | hCoV-19/Denmark/DCGC-1432/2020\|EPI_ISL_622426\|2020 | ···················· | ···················· | ····················· | ···················· | ···················· | ···················· |
| P | 3475 | hCoV-19/Denmark/DCGC-1442/2020\|EPI_ISL_622427\|2020 | ···················· | ···················· | ····················· | ···················· | ···················· | ···················· |
| P | 3476 | hCoV-19/Denmark/DCGC-1446/2020\|EPI_ISL_622428\|2020 | ···················· | ···················· | ····················· | ···················· | ···················· | ···················· |
| P | 3477 | hCoV-19/Denmark/DCGC-1683/2020\|EPI_ISL_622429\|2020 | ···················· | ···················· | ····················· | ···················· | ···················· | ···················· |

| | | | | | | | | |
|---|---|---|---|---|---|---|---|---|
| P | 3478 | hCoV-19/Denmark/DCGC-1390/2020\|EPI_ISL_622430\|2020 | ···················· | ···················· | ···················· | ···················· | ···················· | ···················· |
| P | 3479 | hCoV-19/Denmark/DCGC-1397/2020\|EPI_ISL_622431\|2020 | ···················· | ···················· | ···················· | ···················· | ···················· | ···················· |
| P | 3480 | hCoV-19/Denmark/DCGC-1415/2020\|EPI_ISL_622432\|2020 | ···················· | ···················· | ···················· | ···················· | ···················· | ···················· |
| P | 3481 | hCoV-19/Denmark/DCGC-1427/2020\|EPI_ISL_622433\|2020 | ···················· | ···················· | ···················· | ···················· | ···················· | ···················· |
| P | 3482 | hCoV-19/Denmark/DCGC-1433/2020\|EPI_ISL_622434\|2020 | ···················· | ···················· | ···················· | ···················· | ···················· | ···················· |
| P | 3483 | hCoV-19/Denmark/DCGC-1436/2020\|EPI_ISL_622435\|2020 | ···················· | ···················· | ···················· | ···················· | ···················· | ···················· |
| P | 3484 | hCoV-19/Denmark/DCGC-1443/2020\|EPI_ISL_622436\|2020 | ···················· | ···················· | ···················· | ···················· | ···················· | ···················· |
| P | 3485 | hCoV-19/Denmark/DCGC-1391/2020\|EPI_ISL_622437\|2020 | ···················· | ···················· | ···················· | ···················· | ···················· | ···················· |
| P | 3486 | hCoV-19/Denmark/DCGC-1456/2020\|EPI_ISL_622438\|2020 | ···················· | ···················· | ···················· | ···················· | ···················· | ···················· |
| P | 3487 | hCoV-19/Denmark/DCGC-1409/2020\|EPI_ISL_622439\|2020 | ···················· | ···················· | ···················· | ···················· | ···················· | ···················· |
| P | 3488 | hCoV-19/Denmark/DCGC-1416/2020\|EPI_ISL_622440\|2020 | ···················· | ···················· | ···················· | ···················· | ···················· | ···················· |
| P | 3489 | hCoV-19/Denmark/DCGC-1421/2020\|EPI_ISL_622441\|2020 | ···················· | ···················· | ···················· | ···················· | ···················· | ···················· |
| P | 3490 | hCoV-19/Denmark/DCGC-1428/2020\|EPI_ISL_622442\|2020 | ···················· | ···················· | ···················· | ···················· | ···················· | ···················· |
| P | 3491 | hCoV-19/Denmark/DCGC-1458/2020\|EPI_ISL_622444\|2020 | ···················· | ···················· | ···················· | ···················· | ···················· | ···················· |
| P | 3492 | hCoV-19/Denmark/DCGC-1437/2020\|EPI_ISL_622445\|2020 | ···················· | ···················· | ···················· | ···················· | ···················· | ···················· |
| P | 3493 | hCoV-19/Denmark/DCGC-1444/2020\|EPI_ISL_622446\|2020 | ···················· | ···················· | ···················· | ···················· | ···················· | ···················· |
| P | 3494 | hCoV-19/Denmark/DCGC-1447/2020\|EPI_ISL_622447\|2020 | ···················· | ···················· | ···················· | ···················· | ···················· | ···················· |
| P | 3495 | hCoV-19/Denmark/DCGC-1392/2020\|EPI_ISL_622448\|2020 | ···················· | ···················· | ···················· | ···················· | ···················· | ···················· |
| P | 3496 | hCoV-19/Denmark/DCGC-1405/2020\|EPI_ISL_622450\|2020 | ···················· | ···················· | ···················· | ···················· | ···················· | ···················· |
| P | 3497 | hCoV-19/Denmark/DCGC-1417/2020\|EPI_ISL_622451\|2020 | ···················· | ···················· | ···················· | ···················· | ···················· | ···················· |
| P | 3498 | hCoV-19/Denmark/DCGC-1422/2020\|EPI_ISL_622452\|2020 | ···················· | ···················· | ···················· | ···················· | ···················· | ···················· |
| P | 3499 | hCoV-19/Denmark/DCGC-1434/2020\|EPI_ISL_622453\|2020 | ···················· | ···················· | ···················· | ···················· | ···················· | ···················· |
| P | 3500 | hCoV-19/Denmark/DCGC-1438/2020\|EPI_ISL_622454\|2020 | ···················· | ···················· | ···················· | ···················· | ···················· | ···················· |
| P | 3501 | hCoV-19/Denmark/DCGC-1448/2020\|EPI_ISL_622455\|2020 | ···················· | ···················· | ···················· | ···················· | ···················· | ···················· |
| P | 3502 | hCoV-19/Denmark/DCGC-1393/2020\|EPI_ISL_622456\|2020 | ···················· | ···················· | ···················· | ···················· | ···················· | ···················· |
| P | 3503 | hCoV-19/Denmark/DCGC-1398/2020\|EPI_ISL_622457\|2020 | ···················· | ···················· | ···················· | ···················· | ···················· | ···················· |
| P | 3504 | hCoV-19/Denmark/DCGC-1647/2020\|EPI_ISL_622458\|2020 | ···················· | ···················· | ···················· | ···················· | ···················· | ···················· |
| P | 3505 | hCoV-19/Denmark/DCGC-1406/2020\|EPI_ISL_622459\|2020 | ···················· | ···················· | ···················· | ···················· | ···················· | ···················· |
| P | 3506 | hCoV-19/Denmark/DCGC-1410/2020\|EPI_ISL_622460\|2020 | ···················· | ···················· | ···················· | ···················· | ···················· | ···················· |
| P | 3507 | hCoV-19/Denmark/DCGC-1423/2020\|EPI_ISL_622462\|2020 | ···················· | ···················· | ···················· | ···················· | ···················· | ···················· |
| P | 3508 | hCoV-19/Denmark/DCGC-1429/2020\|EPI_ISL_622463\|2020 | ···················· | ···················· | ···················· | ···················· | ···················· | ···················· |
| P | 3509 | hCoV-19/Denmark/DCGC-1439/2020\|EPI_ISL_622464\|2020 | ···················· | ···················· | ···················· | ···················· | ···················· | ···················· |
| P | 3510 | hCoV-19/Denmark/DCGC-1445/2020\|EPI_ISL_622465\|2020 | ···················· | ···················· | ···················· | ···················· | ···················· | ···················· |
| P | 3511 | hCoV-19/Denmark/DCGC-1394/2020\|EPI_ISL_622467\|2020 | ···················· | ···················· | ···················· | ···················· | ···················· | ···················· |
| P | 3512 | hCoV-19/Denmark/DCGC-1399/2020\|EPI_ISL_622468\|2020 | ···················· | ···················· | ···················· | ···················· | ···················· | ···················· |
| P | 3513 | hCoV-19/Denmark/DCGC-1401/2020\|EPI_ISL_622469\|2020 | ···················· | ···················· | ···················· | ···················· | ···················· | ···················· |
| P | 3514 | hCoV-19/Denmark/DCGC-1457/2020\|EPI_ISL_622470\|2020 | ···················· | ···················· | ···················· | ···················· | ···················· | ···················· |
| P | 3515 | hCoV-19/Denmark/DCGC-1411/2020\|EPI_ISL_622471\|2020 | ···················· | ···················· | ···················· | ···················· | ···················· | ···················· |
| P | 3516 | hCoV-19/Denmark/DCGC-1419/2020\|EPI_ISL_622472\|2020 | ···················· | ···················· | ···················· | ···················· | ···················· | ···················· |
| P | 3517 | hCoV-19/Denmark/DCGC-1424/2020\|EPI_ISL_622473\|2020 | ···················· | ···················· | ···················· | ···················· | ···················· | ···················· |
| P | 3518 | hCoV-19/Denmark/DCGC-1430/2020\|EPI_ISL_622474\|2020 | ···················· | ···················· | ···················· | ···················· | ···················· | ···················· |
| P | 3519 | hCoV-19/Denmark/DCGC-1459/2020\|EPI_ISL_622475\|2020 | ···················· | ···················· | ···················· | ···················· | ···················· | ···················· |
| P | 3520 | hCoV-19/Denmark/DCGC-1440/2020\|EPI_ISL_622476\|2020 | ···················· | ···················· | ···················· | ···················· | ···················· | ···················· |
| P | 3521 | hCoV-19/Denmark/DCGC-1460/2020\|EPI_ISL_622477\|2020 | ···················· | ···················· | ···················· | ···················· | ···················· | ···················· |
| P | 3522 | hCoV-19/Denmark/DCGC-1395/2020\|EPI_ISL_622478\|2020 | ···················· | ···················· | ···················· | ···················· | ···················· | ···················· |
| P | 3523 | hCoV-19/Denmark/DCGC-1400/2020\|EPI_ISL_622479\|2020 | ···················· | ···················· | ···················· | ···················· | ···················· | ···················· |
| P | 3524 | hCoV-19/Denmark/DCGC-1402/2020\|EPI_ISL_622480\|2020 | ···················· | ···················· | ···················· | ···················· | ···················· | ···················· |
| P | 3525 | hCoV-19/Denmark/DCGC-1407/2020\|EPI_ISL_622481\|2020 | ···················· | ···················· | ···················· | ···················· | ···················· | ···················· |
| P | 3526 | hCoV-19/Denmark/DCGC-1412/2020\|EPI_ISL_622482\|2020 | ···················· | ···················· | ···················· | ···················· | ···················· | ···················· |
| P | 3527 | hCoV-19/Denmark/DCGC-1420/2020\|EPI_ISL_622483\|2020 | ···················· | ···················· | ···················· | ···················· | ···················· | ···················· |
| P | 3528 | hCoV-19/Denmark/DCGC-1425/2020\|EPI_ISL_622484\|2020 | ···················· | ···················· | ···················· | ···················· | ···················· | ···················· |
| P | 3529 | hCoV-19/Denmark/DCGC-1431/2020\|EPI_ISL_622485\|2020 | ···················· | ···················· | ···················· | ···················· | ···················· | ···················· |
| P | 3530 | hCoV-19/Denmark/DCGC-1435/2020\|EPI_ISL_622486\|2020 | ···················· | ···················· | ···················· | ···················· | ···················· | ···················· |
| P | 3531 | hCoV-19/Denmark/DCGC-1441/2020\|EPI_ISL_622487\|2020 | ···················· | ···················· | ···················· | ···················· | ···················· | ···················· |
| P | 3532 | hCoV-19/Denmark/DCGC-1461/2020\|EPI_ISL_622488\|2020 | ···················· | ···················· | ···················· | ···················· | ···················· | ···················· |
| P | 3533 | hCoV-19/Denmark/DCGC-1413/2020\|EPI_ISL_622491\|2020 | ···················· | ···················· | ···················· | ···················· | ···················· | ···················· |
| P | 3534 | hCoV-19/Denmark/DCGC-1426/2020\|EPI_ISL_622492\|2020 | ········T··········· | ···················· | ··········T········· | ···················· | ···················· | ···················· |
| P | 3535 | hCoV-19/Denmark/DCGC-1903/2020\|EPI_ISL_622493\|2020 | ···················· | ···················· | ···················· | ···················· | ···················· | ···················· |
| P | 3536 | hCoV-19/Denmark/DCGC-1904/2020\|EPI_ISL_622494\|2020 | ···················· | ···················· | ···················· | ···················· | ···················· | ···················· |
| P | 3537 | hCoV-19/Denmark/DCGC-1905/2020\|EPI_ISL_622495\|2020 | ···················· | ···················· | ···················· | ···················· | ···················· | ···················· |
| P | 3538 | hCoV-19/Denmark/DCGC-1906/2020\|EPI_ISL_622496\|2020 | ···················· | ···················· | ···················· | ···················· | ···················· | ···················· |
| P | 3539 | hCoV-19/Denmark/DCGC-1908/2020\|EPI_ISL_622498\|2020 | ···················· | ···················· | ···················· | ···················· | ···················· | ···················· |
| P | 3540 | hCoV-19/Denmark/DCGC-1909/2020\|EPI_ISL_622499\|2020 | ···················· | ···················· | ···················· | ···················· | ···················· | ···················· |
| P | 3541 | hCoV-19/England/CAMC-AB1D0D/2020\|EPI_ISL_623746\|20 | ···················· | ···················· | ···················· | ···················· | ···················· | ···················· |
| P | 3542 | hCoV-19/England/MILK-AB7DE9/2020\|EPI_ISL_623748\|20 | ···················· | ···················· | ···················· | ···················· | ···················· | ···················· |
| P | 3543 | hCoV-19/England/CAMC-AB21A6/2020\|EPI_ISL_623749\|20 | ···················· | ···················· | ···················· | ···················· | ···················· | ···················· |
| P | 3544 | hCoV-19/England/CAMC-AB1C97/2020\|EPI_ISL_623750\|20 | ···················· | ···················· | ···················· | ···················· | ···················· | ···················· |
| P | 3545 | hCoV-19/England/CAMC-AB1417/2020\|EPI_ISL_623751\|20 | ···················· | ···················· | ···················· | ···················· | ···················· | ···················· |
| P | 3546 | hCoV-19/England/MILK-AB8D7F/2020\|EPI_ISL_623752\|20 | ···················· | ···················· | ···················· | ···················· | ···················· | ···················· |
| P | 3547 | hCoV-19/England/MILK-AB794F/2020\|EPI_ISL_623753\|20 | ···················· | ···················· | ···················· | ···················· | ···················· | ···················· |
| P | 3548 | hCoV-19/England/CAMC-AB1426/2020\|EPI_ISL_623754\|20 | ···················· | ···················· | ···················· | ···················· | ···················· | ···················· |
| P | 3549 | hCoV-19/England/MILK-AB90A2/2020\|EPI_ISL_623755\|20 | ···················· | ···················· | ···················· | ···················· | ···················· | ···················· |
| P | 3550 | hCoV-19/England/MILK-AB8DD9/2020\|EPI_ISL_623756\|20 | ···················· | ···················· | ···················· | ···················· | ···················· | ···················· |
| P | 3551 | hCoV-19/England/MILK-AB7B67/2020\|EPI_ISL_623757\|20 | ···················· | ···················· | ···················· | ···················· | ···················· | ···················· |
| P | 3552 | hCoV-19/England/CAMC-AB1347/2020\|EPI_ISL_623758\|20 | ···················· | ···················· | ···················· | ···················· | ···················· | ···················· |
| P | 3553 | hCoV-19/England/CAMC-AB1B7C/2020\|EPI_ISL_623759\|20 | ···················· | ···················· | ···················· | ···················· | ···················· | ···················· |
| P | 3554 | hCoV-19/England/MILK-AB7BB2/2020\|EPI_ISL_623760\|20 | ···················· | ···················· | ···················· | ···················· | ···················· | ···················· |
| P | 3555 | hCoV-19/England/MILK-AB7B85/2020\|EPI_ISL_623761\|20 | ···················· | ···················· | ···················· | ···················· | ···················· | ···················· |
| P | 3556 | hCoV-19/England/CAMC-AB11A7/2020\|EPI_ISL_623762\|20 | ···················· | ···················· | ···················· | ···················· | ···················· | ···················· |
| P | 3557 | hCoV-19/England/CAMC-AB1D1C/2020\|EPI_ISL_623763\|20 | ···················· | ···················· | ···················· | ···················· | ···················· | ···················· |

| | | | | | | | | | |
|---|---|---|---|---|---|---|---|---|---|
| P | 3558 | hCoV-19/England/CAMC-AB1875/2020\|EPI_ISL_623764\|20 | ···················· | ···················· | ···················· | ···················· | ···················· | ···················· | |
| P | 3559 | hCoV-19/England/MILK-AB7C64/2020\|EPI_ISL_623766\|20 | ···················· | ···················· | ···················· | ···················· | ···················· | ···················· | |
| P | 3560 | hCoV-19/England/MILK-AB751E/2020\|EPI_ISL_623767\|20 | ···················· | ···················· | ···················· | ···················· | ···················· | ···················· | |
| P | 3561 | hCoV-19/England/MILK-AB8DAC/2020\|EPI_ISL_623768\|20 | ···················· | ···················· | ···················· | ···················· | ···················· | ···················· | |
| P | 3562 | hCoV-19/England/MILK-AB8E30/2020\|EPI_ISL_623769\|20 | ···················· | ···················· | ···················· | ···················· | ···················· | ···················· | |
| P | 3563 | hCoV-19/England/MILK-AB9B83/2020\|EPI_ISL_623770\|20 | ···················· | ···················· | ···················· | ···················· | ···················· | ···················· | |
| P | 3564 | hCoV-19/England/MILK-AB93C7/2020\|EPI_ISL_623773\|20 | ···················· | ···················· | ···················· | ···················· | ······T············· | ···············A···· | |
| P | 3565 | hCoV-19/England/CAMC-AB18EE/2020\|EPI_ISL_623774\|20 | ···················· | ···················· | ···················· | ···················· | ···················· | ···················· | |
| P | 3566 | hCoV-19/England/MILK-AB8A2D/2020\|EPI_ISL_623777\|20 | ···················· | ···················· | ···················· | ···················· | ···················· | ···················· | |
| P | 3567 | hCoV-19/England/MILK-AB7D70/2020\|EPI_ISL_623778\|20 | ···················· | ···················· | ···················· | ···················· | ···················· | ···················· | |
| P | 3568 | hCoV-19/England/MILK-AB78E8/2020\|EPI_ISL_623780\|20 | ···················· | ···················· | ···················· | ···················· | ···················· | ···················· | |
| P | 3569 | hCoV-19/England/MILK-AB85B3/2020\|EPI_ISL_623782\|20 | ···················· | ···················· | ···················· | ···················· | ···················· | ···················· | |
| P | 3570 | hCoV-19/England/MILK-AB752D/2020\|EPI_ISL_623783\|20 | ···················· | ···················· | ···················· | ···················· | ···················· | ···················· | |
| P | 3571 | hCoV-19/England/MILK-AB7D25/2020\|EPI_ISL_623784\|20 | ···················· | ···················· | ···················· | ···················· | ···················· | ···················· | |
| P | 3572 | hCoV-19/England/CAMC-AB1DEF/2020\|EPI_ISL_623785\|20 | ···················· | ···················· | ···················· | ···················· | ···················· | ···················· | |
| P | 3573 | hCoV-19/England/MILK-AB90B1/2020\|EPI_ISL_623786\|20 | ···················· | ···················· | ···················· | ···················· | ···················· | ···················· | |
| P | 3574 | hCoV-19/England/MILK-AB7EB9/2020\|EPI_ISL_623787\|20 | ···················· | ···················· | ···················· | ···················· | ···················· | ···················· | |
| P | 3575 | hCoV-19/England/CAMC-AB1356/2020\|EPI_ISL_623788\|20 | ···················· | ···················· | ···················· | ···················· | ···················· | ···················· | |
| P | 3576 | hCoV-19/England/MILK-AB8AB4/2020\|EPI_ISL_623789\|20 | ···················· | ···················· | ···················· | ···················· | ······T············· | ···················· | |
| P | 3577 | hCoV-19/England/CAMC-AB204F/2020\|EPI_ISL_623790\|20 | ···················· | ···················· | ···················· | ···················· | ···················· | ···················· | |
| P | 3578 | hCoV-19/England/CAMC-AB1435/2020\|EPI_ISL_623791\|20 | ···················· | ···················· | ···················· | ···················· | ···················· | ···················· | |
| P | 3579 | hCoV-19/England/MILK-AB7CDD/2020\|EPI_ISL_623792\|20 | ···················· | ···················· | ···················· | ···················· | ···················· | ···················· | |
| P | 3580 | hCoV-19/England/CAMC-AB21D3/2020\|EPI_ISL_623793\|20 | ···················· | ···················· | ···················· | ···················· | ···················· | ···················· | |
| P | 3581 | hCoV-19/England/CAMC-AB1408/2020\|EPI_ISL_623794\|20 | ···················· | ···················· | ···················· | ···················· | ···················· | ···················· | |
| P | 3582 | hCoV-19/England/MILK-AB7903/2020\|EPI_ISL_623795\|20 | ···················· | ···················· | ···················· | ···················· | ···················· | ···················· | |
| P | 3583 | hCoV-19/England/CAMC-AB11C5/2020\|EPI_ISL_623797\|20 | ···················· | ···················· | ···················· | ···················· | ···················· | ···················· | |
| P | 3584 | hCoV-19/England/CAMC-AB1329/2020\|EPI_ISL_623798\|20 | ···················· | ···················· | ···················· | ···················· | ···················· | ···················· | |
| P | 3585 | hCoV-19/Denmark/DCGC-5269/2020\|EPI_ISL_615200\|2020 | ···················· | ···················· | ···················· | ···················· | ···················· | ···················· | |
| P | 3586 | hCoV-19/Denmark/DCGC-5270/2020\|EPI_ISL_615201\|2020 | ···················· | ···················· | ···················· | ···················· | ···················· | ···················· | |
| P | 3587 | hCoV-19/Denmark/DCGC-5271/2020\|EPI_ISL_615202\|2020 | ···················· | ·A·················· | ···················· | ···················· | ···················· | ···A················ | |
| P | 3588 | hCoV-19/Denmark/DCGC-5272/2020\|EPI_ISL_615203\|2020 | ···················· | ···················· | ···················· | ···················· | ···················· | ···················· | |
| P | 3589 | hCoV-19/Denmark/DCGC-5273/2020\|EPI_ISL_615204\|2020 | ···················· | ···················· | ···················· | ···················· | ···················· | ···················· | |
| P | 3590 | hCoV-19/Denmark/DCGC-5274/2020\|EPI_ISL_615205\|2020 | ···················· | ···················· | ···················· | ···················· | ···················· | ···················· | |
| P | 3591 | hCoV-19/Denmark/DCGC-5275/2020\|EPI_ISL_615206\|2020 | ···················· | ···················· | ···················· | ···················· | ···················· | ···················· | |
| P | 3592 | hCoV-19/Denmark/DCGC-5276/2020\|EPI_ISL_615207\|2020 | ···················· | ···················· | ···················· | ···················· | ···················· | ···················· | |
| P | 3593 | hCoV-19/Denmark/DCGC-5277/2020\|EPI_ISL_615208\|2020 | ···················· | ···················· | ···················· | ···················· | ···················· | ···················· | |
| P | 3594 | hCoV-19/Denmark/DCGC-5278/2020\|EPI_ISL_615209\|2020 | ···················· | ···················· | ···················· | ···················· | ···················· | ···················· | |
| P | 3595 | hCoV-19/Denmark/DCGC-5279/2020\|EPI_ISL_615210\|2020 | ···················· | ···················· | ···················· | ···················· | ···················· | ···················· | |
| P | 3596 | hCoV-19/Denmark/DCGC-5280/2020\|EPI_ISL_615211\|2020 | ···················· | ···················· | ···················· | ···················· | ···················· | ···················· | |
| P | 3597 | hCoV-19/Denmark/DCGC-5281/2020\|EPI_ISL_615212\|2020 | ···················· | ···················· | ···················· | ···················· | ···················· | ···················· | |
| P | 3598 | hCoV-19/Denmark/DCGC-5282/2020\|EPI_ISL_615213\|2020 | ···················· | ···················· | ···················· | ···················· | ···················· | ···················· | |
| P | 3599 | hCoV-19/Denmark/DCGC-5283/2020\|EPI_ISL_615214\|2020 | ···················· | ···················· | ···················· | ···················· | ···················· | ···················· | |
| P | 3600 | hCoV-19/Denmark/DCGC-5284/2020\|EPI_ISL_615215\|2020 | ···················· | ···················· | ···················· | ···················· | ···················· | ···················· | |
| P | 3601 | hCoV-19/Denmark/DCGC-5285/2020\|EPI_ISL_615216\|2020 | ···················· | ···················· | ···················· | ···················· | ···················· | ···················· | |
| P | 3602 | hCoV-19/Denmark/DCGC-5286/2020\|EPI_ISL_615217\|2020 | ···················· | ···················· | ···················· | ···················· | ···················· | ···················· | |
| P | 3603 | hCoV-19/Denmark/DCGC-5289/2020\|EPI_ISL_615220\|2020 | ···················· | ···················· | ···················· | ···················· | ···················· | ···················· | |
| P | 3604 | hCoV-19/Denmark/DCGC-5290/2020\|EPI_ISL_615221\|2020 | ···················· | ···················· | ···················· | ···················· | ···················· | ···················· | |
| P | 3605 | hCoV-19/Denmark/DCGC-5291/2020\|EPI_ISL_615222\|2020 | ··················T· | ···················· | ···················· | ···················· | ···················· | ···················· | 1 |
| P | 3606 | hCoV-19/Denmark/DCGC-5292/2020\|EPI_ISL_615223\|2020 | ···················· | ···················· | ···················· | ···················· | ···················· | ···················· | |
| P | 3607 | hCoV-19/Denmark/DCGC-5293/2020\|EPI_ISL_615224\|2020 | ···················· | ···················· | ···················· | ···················· | ···················· | ···················· | |
| P | 3608 | hCoV-19/Denmark/DCGC-5294/2020\|EPI_ISL_615225\|2020 | ···················· | ···················· | ···················· | ···················· | ···················· | ···················· | |
| P | 3609 | hCoV-19/Denmark/DCGC-5295/2020\|EPI_ISL_615226\|2020 | ···················· | ···················· | ···················· | ···················· | ···················· | ···················· | |
| P | 3610 | hCoV-19/Denmark/DCGC-5296/2020\|EPI_ISL_615227\|2020 | ···················· | ···················· | ···················· | ···················· | ···················· | ···················· | |
| P | 3611 | hCoV-19/Denmark/DCGC-5297/2020\|EPI_ISL_615228\|2020 | ···················· | ···················· | ···················· | ···················· | ···················· | ···················· | |
| P | 3612 | hCoV-19/Denmark/DCGC-5298/2020\|EPI_ISL_615229\|2020 | ···················· | ···················· | ···················· | ···················· | ···················· | ···················· | |
| P | 3613 | hCoV-19/Denmark/DCGC-5299/2020\|EPI_ISL_615230\|2020 | ···················· | ···················· | ···················· | ···················· | ···················· | ···················· | |
| P | 3614 | hCoV-19/Denmark/DCGC-5301/2020\|EPI_ISL_615232\|2020 | ···················· | ···················· | ···················· | ···················· | ···················· | ···················· | |
| P | 3615 | hCoV-19/Denmark/DCGC-5302/2020\|EPI_ISL_615233\|2020 | ···················· | ···················· | ···················· | ···················· | ···················· | ···················· | |
| P | 3616 | hCoV-19/Denmark/DCGC-5303/2020\|EPI_ISL_615234\|2020 | ···················· | ···················· | ···················· | ···················· | ···················· | ···················· | |
| P | 3617 | hCoV-19/Denmark/DCGC-5304/2020\|EPI_ISL_615235\|2020 | ···················· | ···················· | ···················· | ···················· | ···················· | ···················· | |
| P | 3618 | hCoV-19/Denmark/DCGC-5305/2020\|EPI_ISL_615236\|2020 | ···················· | ···················· | ···················· | ···················· | ···················· | ···················· | |
| P | 3619 | hCoV-19/Denmark/DCGC-5306/2020\|EPI_ISL_615237\|2020 | ···················· | ···················· | ···················· | ···················· | ···················· | ···················· | |
| P | 3620 | hCoV-19/Denmark/DCGC-5307/2020\|EPI_ISL_615238\|2020 | ····N··············· | ···················· | ···················· | ···················· | ···················· | ···················· | |
| P | 3621 | hCoV-19/Denmark/DCGC-5308/2020\|EPI_ISL_615239\|2020 | ···················· | ···················· | ···················· | ···················· | ···················· | ···················· | |
| P | 3622 | hCoV-19/Denmark/DCGC-5319/2020\|EPI_ISL_615250\|2020 | ···················· | ···················· | ···················· | ···················· | ···················· | ···················· | |
| P | 3623 | hCoV-19/Denmark/DCGC-5320/2020\|EPI_ISL_615251\|2020 | ···················· | ···················· | ···················· | ···················· | ···················· | ···················· | |
| P | 3624 | hCoV-19/Denmark/DCGC-5321/2020\|EPI_ISL_615252\|2020 | ···················· | ···················· | ···················· | ···················· | ···················· | ···················· | |
| P | 3625 | hCoV-19/Denmark/DCGC-5322/2020\|EPI_ISL_615253\|2020 | ···················· | ···················· | ···················· | ···················· | ···················· | ···················· | |
| P | 3626 | hCoV-19/Denmark/DCGC-5323/2020\|EPI_ISL_615254\|2020 | ···················· | ···················· | ···················· | ···················· | ···················· | ···················· | |
| P | 3627 | hCoV-19/Denmark/DCGC-5324/2020\|EPI_ISL_615255\|2020 | ···················· | ···················· | ···················· | ···················· | ···················· | ···················· | |
| P | 3628 | hCoV-19/Denmark/DCGC-5325/2020\|EPI_ISL_615256\|2020 | ···················· | ···················· | ···················· | ···················· | ···················· | ···················· | |
| P | 3629 | hCoV-19/Denmark/DCGC-5326/2020\|EPI_ISL_615257\|2020 | ···················· | ···················· | ···················· | ···················· | ···················· | ···················· | |
| P | 3630 | hCoV-19/Denmark/DCGC-5327/2020\|EPI_ISL_615258\|2020 | ··················T· | ···················· | ···················· | ···················· | ···················· | ···················· | 1 |
| P | 3631 | hCoV-19/Denmark/DCGC-5328/2020\|EPI_ISL_615259\|2020 | ···················· | ·A·················· | ···················· | ···················· | ···················· | ···················· | |
| P | 3632 | hCoV-19/Denmark/DCGC-5329/2020\|EPI_ISL_615260\|2020 | ···················· | ···················· | ···················· | ···················· | ···················· | ···················· | |
| P | 3633 | hCoV-19/Denmark/DCGC-5330/2020\|EPI_ISL_615261\|2020 | ···················· | ···················· | ···················· | ···················· | ···················· | ···················· | |
| P | 3634 | hCoV-19/Denmark/DCGC-5331/2020\|EPI_ISL_615262\|2020 | ···················· | ···················· | ···················· | ···················· | ···················· | ···················· | |
| P | 3635 | hCoV-19/Denmark/DCGC-5333/2020\|EPI_ISL_615264\|2020 | ···················· | ···················· | ···················· | ···················· | ···················· | ···················· | |
| P | 3636 | hCoV-19/Denmark/DCGC-5334/2020\|EPI_ISL_615265\|2020 | ···················· | ···················· | ···················· | ···················· | ···················· | ···················· | |
| P | 3637 | hCoV-19/Denmark/DCGC-5335/2020\|EPI_ISL_615266\|2020 | ···················· | ···················· | ···················· | ···················· | ···················· | ···················· | |

| | | | | | | | | | |
|---|---|---|---|---|---|---|---|---|---|
| P | 3638 | hCoV-19/Denmark/DCGC-5336/2020\|EPI_ISL_615267\|2020 | ···················· | ···················· | ···················· | ···················· | ···················· | ···················· | |
| P | 3639 | hCoV-19/Denmark/DCGC-5337/2020\|EPI_ISL_615268\|2020 | ···················· | ···················· | ···················· | ···················· | ···················· | ···················· | |
| P | 3640 | hCoV-19/Denmark/DCGC-5338/2020\|EPI_ISL_615269\|2020 | ···················· | ···················· | ···················· | ···················· | ···················· | ···················· | |
| P | 3641 | hCoV-19/Denmark/DCGC-5339/2020\|EPI_ISL_615270\|2020 | ···················· | ···················· | ···················· | ···················· | ···················· | ···················· | |
| P | 3642 | hCoV-19/Denmark/DCGC-5341/2020\|EPI_ISL_615272\|2020 | ··················T· | ···················· | ···················· | ···················· | ···················· | ···················· | 1 |
| P | 3643 | hCoV-19/Denmark/DCGC-5342/2020\|EPI_ISL_615273\|2020 | ···················· | ···················· | ···················· | ···················· | ···················· | ···················· | |
| P | 3644 | hCoV-19/Denmark/DCGC-5343/2020\|EPI_ISL_615274\|2020 | ···················· | ···················· | ···················· | ···················· | ···················· | ···················· | |
| P | 3645 | hCoV-19/Denmark/DCGC-5344/2020\|EPI_ISL_615275\|2020 | ···················· | ···················· | ···················· | ···················· | ···················· | ···················· | |
| P | 3646 | hCoV-19/Denmark/DCGC-5345/2020\|EPI_ISL_615276\|2020 | ···················· | ····G··············· | ···················· | ···················· | ···················· | ···················· | |
| P | 3647 | hCoV-19/Denmark/DCGC-5346/2020\|EPI_ISL_615277\|2020 | ···················· | ···················· | ···················· | ···················· | ···················· | ···················· | |
| P | 3648 | hCoV-19/Denmark/DCGC-5347/2020\|EPI_ISL_615278\|2020 | ···················· | ···················· | ···················· | ···················· | ···················· | ···················· | |
| P | 3649 | hCoV-19/Denmark/DCGC-5348/2020\|EPI_ISL_615279\|2020 | ···················· | ···················· | ···················· | ···················· | ···················· | ···················· | |
| P | 3650 | hCoV-19/Denmark/DCGC-5349/2020\|EPI_ISL_615280\|2020 | ···················· | ···················· | ···················· | ···················· | ···················· | ···················· | |
| P | 3651 | hCoV-19/Denmark/DCGC-5350/2020\|EPI_ISL_615281\|2020 | ···················· | ···················· | ···················· | ···················· | ···················· | ···················· | |
| P | 3652 | hCoV-19/Denmark/DCGC-5351/2020\|EPI_ISL_615282\|2020 | ···················· | ···················· | ···················· | ···················· | ···················· | ···················· | |
| P | 3653 | hCoV-19/Denmark/DCGC-5352/2020\|EPI_ISL_615283\|2020 | ···················· | ···················· | ···················· | ···················· | ···················· | ···················· | |
| P | 3654 | hCoV-19/Denmark/DCGC-5353/2020\|EPI_ISL_615284\|2020 | ···················· | ·A·················· | ···················· | ···················· | ···················· | ···A················ | |
| P | 3655 | hCoV-19/Denmark/DCGC-5354/2020\|EPI_ISL_615285\|2020 | ···················· | ···················· | ···················· | ···················· | ···················· | ···················· | |
| P | 3656 | hCoV-19/Denmark/DCGC-5355/2020\|EPI_ISL_615286\|2020 | ···················· | ···················· | ···················· | ···················· | ···················· | ···················· | |
| P | 3657 | hCoV-19/Denmark/DCGC-5356/2020\|EPI_ISL_615287\|2020 | ···················· | ···················· | ···················· | ···················· | ···················· | ···················· | |
| P | 3658 | hCoV-19/Denmark/DCGC-5357/2020\|EPI_ISL_615288\|2020 | ···················· | ···················· | ···················· | ···················· | ···················· | ···················· | |
| P | 3659 | hCoV-19/Denmark/DCGC-5358/2020\|EPI_ISL_615289\|2020 | ···················· | ···················· | ···················· | ···················· | ···················· | ···················· | |
| P | 3660 | hCoV-19/Denmark/DCGC-2543/2020\|EPI_ISL_616519\|2020 | ···················· | ···················· | ···················· | ···················· | ······T············· | ···················· | |
| P | 3661 | hCoV-19/Denmark/DCGC-2544/2020\|EPI_ISL_616520\|2020 | ···················· | ···················· | ···················· | ···················· | ···················· | ···················· | |
| P | 3662 | hCoV-19/Denmark/DCGC-2545/2020\|EPI_ISL_616521\|2020 | ···················· | ···················· | ···················· | ···················· | ···················· | ···················· | |
| P | 3663 | hCoV-19/Denmark/DCGC-2550/2020\|EPI_ISL_616526\|2020 | ···················· | ···················· | ···················· | ···················· | ···················· | ···················· | |
| P | 3664 | hCoV-19/Denmark/DCGC-2551/2020\|EPI_ISL_616527\|2020 | ···················· | ···················· | ···················· | ···················· | ···················· | ···················· | |
| P | 3665 | hCoV-19/Denmark/DCGC-2552/2020\|EPI_ISL_616528\|2020 | ···················· | ···················· | ···················· | ···················· | ···················· | ···················· | |
| P | 3666 | hCoV-19/Denmark/DCGC-2554/2020\|EPI_ISL_616530\|2020 | ···················· | ···················· | ···················· | ···················· | ···················· | ···················· | |
| P | 3667 | hCoV-19/Denmark/DCGC-2555/2020\|EPI_ISL_616531\|2020 | ···················· | ···················· | ···················· | ···················· | ···················· | ···················· | |
| P | 3668 | hCoV-19/Denmark/DCGC-2556/2020\|EPI_ISL_616532\|2020 | ···················· | ···················· | ···················· | ···················· | ···················· | ···················· | |
| P | 3669 | hCoV-19/Denmark/DCGC-2900/2020\|EPI_ISL_616534\|2020 | ···················· | ···················· | ···················· | ···················· | ···················· | ···················· | |
| P | 3670 | hCoV-19/Denmark/DCGC-2901/2020\|EPI_ISL_616535\|2020 | ···················· | ···················· | ···················· | ···················· | ···················· | ···················· | |
| P | 3671 | hCoV-19/Denmark/DCGC-2902/2020\|EPI_ISL_616536\|2020 | ···················· | ···················· | ···················· | ···················· | ···················· | ···················· | |
| P | 3672 | hCoV-19/Denmark/DCGC-2903/2020\|EPI_ISL_616537\|2020 | ···················· | ···················· | ···················· | ···················· | ···················· | ···················· | |
| P | 3673 | hCoV-19/Denmark/DCGC-2904/2020\|EPI_ISL_616538\|2020 | ···················· | ···················· | ···················· | ···················· | ······T············· | ···················· | |
| P | 3674 | hCoV-19/Denmark/DCGC-2906/2020\|EPI_ISL_616540\|2020 | ···················· | ···················· | ···················· | ···················· | ···················· | ···················· | |
| P | 3675 | hCoV-19/Denmark/DCGC-2907/2020\|EPI_ISL_616541\|2020 | ···················· | ···················· | ···················· | ···················· | ···················· | ···················· | |
| P | 3676 | hCoV-19/Denmark/DCGC-2908/2020\|EPI_ISL_616542\|2020 | ···················· | ···················· | ···················· | ···················· | ···················· | ···················· | |
| P | 3677 | hCoV-19/Denmark/DCGC-2909/2020\|EPI_ISL_616543\|2020 | ···················· | ···················· | ···················· | ···················· | ···················· | ···················· | |
| P | 3678 | hCoV-19/Denmark/DCGC-2911/2020\|EPI_ISL_616545\|2020 | ···················· | ···················· | ···················· | ···················· | ···················· | ···················· | |
| P | 3679 | hCoV-19/Denmark/DCGC-2912/2020\|EPI_ISL_616546\|2020 | ···················· | ···················· | ···················· | ···················· | ···················· | ···················· | |
| P | 3680 | hCoV-19/Denmark/DCGC-2913/2020\|EPI_ISL_616547\|2020 | ···················· | ···················· | ···················· | ···················· | ···················· | ···················· | |
| P | 3681 | hCoV-19/Denmark/DCGC-2924/2020\|EPI_ISL_616548\|2020 | ···················· | ···················· | ···················· | ···················· | ···················· | ···················· | |
| P | 3682 | hCoV-19/Denmark/DCGC-2914/2020\|EPI_ISL_616549\|2020 | ···················· | ···················· | ···················· | ···················· | ···················· | ···················· | |
| P | 3683 | hCoV-19/Denmark/DCGC-2915/2020\|EPI_ISL_616550\|2020 | ···················· | ···················· | ···················· | ···················· | ···················· | ···················· | |
| P | 3684 | hCoV-19/Denmark/DCGC-2916/2020\|EPI_ISL_616551\|2020 | ···················· | ···················· | ···················· | ···················· | ···················· | ···················· | |
| P | 3685 | hCoV-19/Denmark/DCGC-2917/2020\|EPI_ISL_616552\|2020 | ···················· | ···················· | ···················· | ···················· | ···················· | ···················· | |
| P | 3686 | hCoV-19/Denmark/DCGC-2918/2020\|EPI_ISL_616553\|2020 | ···················· | ···················· | ···················· | ···················· | ···················· | ···················· | |
| P | 3687 | hCoV-19/Denmark/DCGC-2919/2020\|EPI_ISL_616554\|2020 | ···················· | ···················· | ···················· | ···················· | ···················· | ···················· | |
| P | 3688 | hCoV-19/Denmark/DCGC-2920/2020\|EPI_ISL_616555\|2020 | ···················· | ···················· | ···················· | ···················· | ···················· | ···················· | |
| P | 3689 | hCoV-19/Denmark/DCGC-2921/2020\|EPI_ISL_616556\|2020 | ···················· | ···················· | ···················· | ···················· | ···················· | ···················· | |
| P | 3690 | hCoV-19/Denmark/DCGC-2922/2020\|EPI_ISL_616557\|2020 | ···················· | ···················· | ···················· | ···················· | ···················· | ···················· | |
| P | 3691 | hCoV-19/Denmark/DCGC-2923/2020\|EPI_ISL_616558\|2020 | ···················· | ···················· | ···················· | ···················· | ···················· | ···················· | |
| P | 3692 | hCoV-19/Denmark/DCGC-2865/2020\|EPI_ISL_616559\|2020 | ···················· | ···················· | ···················· | ···················· | ···················· | ···················· | |
| P | 3693 | hCoV-19/Denmark/DCGC-2866/2020\|EPI_ISL_616560\|2020 | ···················· | ···················· | ···················· | ···················· | ···················· | ···················· | |
| P | 3694 | hCoV-19/Denmark/DCGC-2867/2020\|EPI_ISL_616561\|2020 | ···················· | ···················· | ···················· | ···················· | ···················· | ···················· | |
| P | 3695 | hCoV-19/Denmark/DCGC-2869/2020\|EPI_ISL_616563\|2020 | ···················· | ···················· | ···················· | ···················· | ···················· | ···················· | |
| P | 3696 | hCoV-19/Denmark/DCGC-2870/2020\|EPI_ISL_616564\|2020 | ···················· | ···················· | ···················· | ···················· | ···················· | ···················· | |
| P | 3697 | hCoV-19/Denmark/DCGC-2871/2020\|EPI_ISL_616565\|2020 | ···················· | ···················· | ···················· | ···················· | ···················· | ···················· | |
| P | 3698 | hCoV-19/Denmark/DCGC-2872/2020\|EPI_ISL_616566\|2020 | ···················· | ···················· | ···················· | ···················· | ···················· | ···················· | |
| P | 3699 | hCoV-19/Denmark/DCGC-2873/2020\|EPI_ISL_616567\|2020 | ···················· | ···················· | ···················· | ···················· | ···················· | ···················· | |
| P | 3700 | hCoV-19/Denmark/DCGC-2874/2020\|EPI_ISL_616568\|2020 | ···················· | ···················· | ···················· | ···················· | ···················· | ···················· | |
| P | 3701 | hCoV-19/Denmark/DCGC-1715/2020\|EPI_ISL_616569\|2020 | ···················· | ···················· | ···················· | ···················· | ···················· | ···················· | |
| P | 3702 | hCoV-19/Denmark/DCGC-2885/2020\|EPI_ISL_616580\|2020 | ···················· | ···················· | ···················· | ···················· | ···················· | ···················· | |
| P | 3703 | hCoV-19/Denmark/DCGC-2886/2020\|EPI_ISL_616581\|2020 | ···················· | ···················· | ···················· | ···················· | ···················· | ···················· | |
| P | 3704 | hCoV-19/Denmark/DCGC-2887/2020\|EPI_ISL_616582\|2020 | ···················· | ····A··············· | ···················· | ···················· | ···················· | ···················· | |
| P | 3705 | hCoV-19/Denmark/DCGC-2890/2020\|EPI_ISL_616585\|2020 | ···················· | ···················· | ···················· | ···················· | ···················· | ···················· | |
| P | 3706 | hCoV-19/Denmark/DCGC-2891/2020\|EPI_ISL_616586\|2020 | ···················· | ·A·················· | ···················· | ···················· | ···················· | ···A················ | |
| P | 3707 | hCoV-19/Denmark/DCGC-2892/2020\|EPI_ISL_616587\|2020 | ···················· | ···················· | ···················· | ···················· | ···················· | ···················· | |
| P | 3708 | hCoV-19/Denmark/DCGC-2893/2020\|EPI_ISL_616588\|2020 | ···················· | ·A·················· | ···················· | ···················· | ···················· | ···A················ | |
| P | 3709 | hCoV-19/Denmark/DCGC-2755/2020\|EPI_ISL_616589\|2020 | ···················· | ···················· | ···················· | ···················· | ···················· | ···················· | |
| P | 3710 | hCoV-19/Denmark/DCGC-2756/2020\|EPI_ISL_616590\|2020 | ···················· | ···················· | ···················· | ···················· | ···················· | ···················· | |
| P | 3711 | hCoV-19/Denmark/DCGC-2757/2020\|EPI_ISL_616591\|2020 | ···················· | ···················· | ···················· | ···················· | ···················· | ···················· | |
| P | 3712 | hCoV-19/Denmark/DCGC-2758/2020\|EPI_ISL_616592\|2020 | ···················· | ···················· | ···················· | ···················· | ···················· | ···················· | |
| P | 3713 | hCoV-19/Denmark/DCGC-2759/2020\|EPI_ISL_616593\|2020 | ···················· | ···················· | ···················· | ···················· | ···················· | ···················· | |
| P | 3714 | hCoV-19/Denmark/DCGC-2760/2020\|EPI_ISL_616594\|2020 | ···················· | ···················· | ···················· | ···················· | ······T············· | ···················· | |
| P | 3715 | hCoV-19/Denmark/DCGC-2761/2020\|EPI_ISL_616595\|2020 | ···················· | ···················· | ···················· | ···················· | ···················· | ···················· | |
| P | 3716 | hCoV-19/Denmark/DCGC-2762/2020\|EPI_ISL_616596\|2020 | ···················· | ···················· | ···················· | ···················· | ······T············· | ···················· | |
| P | 3717 | hCoV-19/Denmark/DCGC-2763/2020\|EPI_ISL_616597\|2020 | ···················· | ···················· | ···················· | ···················· | ······T············· | ···················· | |

| | | | | | | | | |
|---|---|---|---|---|---|---|---|---|
| P | 3718 | hCoV-19/Denmark/DCGC-2764/2020\|EPI_ISL_616598\|2020 | .................... | .................... | .................... | .................... | .................... | .................... |
| P | 3719 | hCoV-19/Denmark/DCGC-2765/2020\|EPI_ISL_616599\|2020 | .................... | .A.................. | .................... | .................... | .................... | .................... |
| P | 3720 | hCoV-19/Denmark/DCGC-2530/2020\|EPI_ISL_617800\|2020 | .................... | .................... | .................... | .................... | .................... | .................... |
| P | 3721 | hCoV-19/Denmark/DCGC-2531/2020\|EPI_ISL_617801\|2020 | .................... | .................... | .................... | .................... | .................... | .................... |
| P | 3722 | hCoV-19/Denmark/DCGC-2567/2020\|EPI_ISL_617802\|2020 | .................... | .................... | .................... | .................... | .................... | .................... |
| P | 3723 | hCoV-19/Denmark/DCGC-2532/2020\|EPI_ISL_617803\|2020 | .................... | .................... | .................... | .................... | .................... | .................... |
| P | 3724 | hCoV-19/Denmark/DCGC-2533/2020\|EPI_ISL_617804\|2020 | .................... | .................... | .................... | .................... | .................... | .................... |
| P | 3725 | hCoV-19/Denmark/DCGC-2534/2020\|EPI_ISL_617805\|2020 | .................... | .................... | .................... | .................... | .................... | .................... |
| P | 3726 | hCoV-19/Denmark/DCGC-2535/2020\|EPI_ISL_617806\|2020 | .................... | .................... | .................... | .................... | .................... | .................... |
| P | 3727 | hCoV-19/Denmark/DCGC-2536/2020\|EPI_ISL_617807\|2020 | .................... | .................... | .................... | .................... | .................... | .................... |
| P | 3728 | hCoV-19/Denmark/DCGC-2537/2020\|EPI_ISL_617808\|2020 | .................... | .................... | .................... | .................... | .................... | .................... |
| P | 3729 | hCoV-19/Denmark/DCGC-2538/2020\|EPI_ISL_617809\|2020 | .................... | .................... | .................... | .................... | .................... | .................... |
| P | 3730 | hCoV-19/Denmark/DCGC-2539/2020\|EPI_ISL_617810\|2020 | .................... | .................... | .................... | .................... | .................... | .................... |
| P | 3731 | hCoV-19/Denmark/DCGC-2540/2020\|EPI_ISL_617811\|2020 | .................... | .................... | .................... | .................... | .................... | .................... |
| P | 3732 | hCoV-19/Denmark/DCGC-2541/2020\|EPI_ISL_617812\|2020 | .................... | .................... | .................... | .................... | .................... | .................... |
| P | 3733 | hCoV-19/Denmark/DCGC-2568/2020\|EPI_ISL_617813\|2020 | .................... | .................... | .................... | .................... | .................... | .................... |
| P | 3734 | hCoV-19/Denmark/DCGC-1712/2020\|EPI_ISL_617814\|2020 | .................... | .................... | .................... | .................... | .................... | .................... |
| P | 3735 | hCoV-19/Denmark/DCGC-1711/2020\|EPI_ISL_617815\|2020 | .................... | .................... | .................... | .................... | .................... | .................... |
| P | 3736 | hCoV-19/Denmark/DCGC-2558/2020\|EPI_ISL_617816\|2020 | .................... | .................... | .................... | .................... | .................... | .................... |
| P | 3737 | hCoV-19/Denmark/DCGC-2559/2020\|EPI_ISL_617817\|2020 | .................... | .................... | .................... | .................... | .................... | .................... |
| P | 3738 | hCoV-19/Denmark/DCGC-2560/2020\|EPI_ISL_617818\|2020 | .................... | .................... | .................... | .................... | .................... | .................... |
| P | 3739 | hCoV-19/Denmark/DCGC-2561/2020\|EPI_ISL_617819\|2020 | .................... | .................... | .................... | .................... | .................... | .................... |
| P | 3740 | hCoV-19/Denmark/DCGC-2562/2020\|EPI_ISL_617820\|2020 | .................... | .................... | .................... | .................... | .................... | .................... |
| P | 3741 | hCoV-19/Denmark/DCGC-2563/2020\|EPI_ISL_617821\|2020 | .................... | .................... | .................... | .................... | .................... | .................... |
| P | 3742 | hCoV-19/Denmark/DCGC-2564/2020\|EPI_ISL_617822\|2020 | .................... | .................... | .................... | .................... | .................... | .................... |
| P | 3743 | hCoV-19/Denmark/DCGC-2587/2020\|EPI_ISL_617823\|2020 | .................... | .................... | .................... | .................... | .................... | .................... |
| P | 3744 | hCoV-19/Denmark/DCGC-2588/2020\|EPI_ISL_617824\|2020 | .................... | .................... | .................... | .................... | .................... | .................... |
| P | 3745 | hCoV-19/Denmark/DCGC-2589/2020\|EPI_ISL_617825\|2020 | .................... | .................... | .................... | .................... | .................... | .................... |
| P | 3746 | hCoV-19/Denmark/DCGC-2590/2020\|EPI_ISL_617826\|2020 | .................... | .A.................. | .................... | .................... | .................... | .................... |
| P | 3747 | hCoV-19/Denmark/DCGC-2591/2020\|EPI_ISL_617827\|2020 | .................... | .A.................. | .................... | .................... | .................... | .................... |
| P | 3748 | hCoV-19/Denmark/DCGC-2614/2020\|EPI_ISL_617851\|2020 | .................... | .................... | .................... | .................... | .................... | .................... |
| P | 3749 | hCoV-19/Denmark/DCGC-2615/2020\|EPI_ISL_617852\|2020 | .................... | .................... | .................... | .................... | .................... | .................... |
| P | 3750 | hCoV-19/Denmark/DCGC-2618/2020\|EPI_ISL_617855\|2020 | .................... | .................... | .................... | .................... | .................... | .................... |
| P | 3751 | hCoV-19/Denmark/DCGC-2619/2020\|EPI_ISL_617856\|2020 | .................... | .................... | .................... | .................... | .................... | .................... |
| P | 3752 | hCoV-19/Denmark/DCGC-2641/2020\|EPI_ISL_617860\|2020 | .................... | .................... | .................... | .................... | .................... | .................... |
| P | 3753 | hCoV-19/Denmark/DCGC-2642/2020\|EPI_ISL_617861\|2020 | .................... | .................... | .................... | .................... | .................... | .................... |
| P | 3754 | hCoV-19/Denmark/DCGC-2643/2020\|EPI_ISL_617862\|2020 | .................... | .A.................. | .................... | .................... | .................... | .................... |
| P | 3755 | hCoV-19/Denmark/DCGC-2644/2020\|EPI_ISL_617863\|2020 | .................... | .................... | .................... | .................... | .................... | .................... |
| P | 3756 | hCoV-19/Denmark/DCGC-2645/2020\|EPI_ISL_617864\|2020 | .................... | .................... | .................... | .................... | .................... | .................... |
| P | 3757 | hCoV-19/Denmark/DCGC-2646/2020\|EPI_ISL_617865\|2020 | .................... | .................... | .................... | .................... | .................... | .................... |
| P | 3758 | hCoV-19/Denmark/DCGC-2647/2020\|EPI_ISL_617866\|2020 | .................... | .................... | .................... | .................... | .................... | .................... |
| P | 3759 | hCoV-19/Denmark/DCGC-2649/2020\|EPI_ISL_617868\|2020 | .................... | .................... | .................... | .................... | .................... | .................... |
| P | 3760 | hCoV-19/Denmark/DCGC-2570/2020\|EPI_ISL_617869\|2020 | .................... | .................... | .................... | .................... | .................... | .................... |
| P | 3761 | hCoV-19/Denmark/DCGC-2651/2020\|EPI_ISL_617871\|2020 | .................... | .................... | .................... | .................... | .................... | .................... |
| P | 3762 | hCoV-19/Denmark/DCGC-2652/2020\|EPI_ISL_617872\|2020 | .................... | .................... | .................... | .................... | .................... | .................... |
| P | 3763 | hCoV-19/Denmark/DCGC-2653/2020\|EPI_ISL_617873\|2020 | .................... | .................... | .................... | .................... | .................... | .................... |
| P | 3764 | hCoV-19/Denmark/DCGC-2654/2020\|EPI_ISL_617874\|2020 | .................... | .................... | .................... | .................... | .................... | .................... |
| P | 3765 | hCoV-19/Denmark/DCGC-2655/2020\|EPI_ISL_617875\|2020 | .................... | .................... | .................... | .................... | .................... | .................... |
| P | 3766 | hCoV-19/Denmark/DCGC-2656/2020\|EPI_ISL_617876\|2020 | .................... | .................... | .................... | .................... | .................... | .................... |
| P | 3767 | hCoV-19/Denmark/DCGC-2657/2020\|EPI_ISL_617877\|2020 | .................... | .................... | .................... | .................... | .................... | .................... |
| P | 3768 | hCoV-19/Denmark/DCGC-2659/2020\|EPI_ISL_617879\|2020 | .................... | .................... | .................... | .................... | .................... | .................... |
| P | 3769 | hCoV-19/Denmark/DCGC-2661/2020\|EPI_ISL_617881\|2020 | .................... | .................... | .................... | .................... | .................... | .................... |
| P | 3770 | hCoV-19/Denmark/DCGC-2662/2020\|EPI_ISL_617882\|2020 | .................... | .................... | .................... | .................... | .................... | .................... |
| P | 3771 | hCoV-19/Denmark/DCGC-2663/2020\|EPI_ISL_617883\|2020 | .................... | .................... | .................... | .................... | .................... | .................... |
| P | 3772 | hCoV-19/Denmark/DCGC-2664/2020\|EPI_ISL_617884\|2020 | .................... | .................... | .................... | .................... | .................... | .................... |
| P | 3773 | hCoV-19/Denmark/DCGC-2665/2020\|EPI_ISL_617885\|2020 | .................... | .................... | .................... | .................... | .................... | .................... |
| P | 3774 | hCoV-19/Denmark/DCGC-2666/2020\|EPI_ISL_617886\|2020 | .................... | .................... | .................... | .................... | .................... | .................... |
| P | 3775 | hCoV-19/Denmark/DCGC-2667/2020\|EPI_ISL_617887\|2020 | .................... | .A.................. | .................... | .................... | .................... | .................... |
| P | 3776 | hCoV-19/Denmark/DCGC-2668/2020\|EPI_ISL_617888\|2020 | .................... | .................... | .................... | .................... | .................... | .................... |
| P | 3777 | hCoV-19/Denmark/DCGC-2669/2020\|EPI_ISL_617889\|2020 | .................... | .................... | .................... | .................... | .................... | .................... |
| P | 3778 | hCoV-19/Denmark/DCGC-2670/2020\|EPI_ISL_617890\|2020 | .................... | .................... | .................... | .................... | .................... | .................... |
| P | 3779 | hCoV-19/Denmark/DCGC-2671/2020\|EPI_ISL_617891\|2020 | .................... | .................... | .................... | .................... | .................... | .................... |
| P | 3780 | hCoV-19/Denmark/DCGC-2673/2020\|EPI_ISL_617893\|2020 | .................... | .................... | .................... | .................... | .................... | .................... |
| P | 3781 | hCoV-19/Denmark/DCGC-2571/2020\|EPI_ISL_617894\|2020 | .................... | .................... | .................... | .................... | .................... | .................... |
| P | 3782 | hCoV-19/Denmark/DCGC-2674/2020\|EPI_ISL_617895\|2020 | .................... | .................... | .................... | .................... | .................... | .................... |
| P | 3783 | hCoV-19/Denmark/DCGC-2675/2020\|EPI_ISL_617896\|2020 | .................... | .................... | .................... | .................... | .................... | .................... |
| P | 3784 | hCoV-19/Denmark/DCGC-2676/2020\|EPI_ISL_617897\|2020 | .................... | .................... | .................... | .................... | .................... | .................... |
| P | 3785 | hCoV-19/Denmark/DCGC-2677/2020\|EPI_ISL_617898\|2020 | .................... | .................... | .................... | .................... | .................... | .................... |
| P | 3786 | hCoV-19/Denmark/DCGC-2678/2020\|EPI_ISL_617899\|2020 | .................... | .................... | .................... | .................... | .................... | .................... |
| P | 3787 | hCoV-19/Denmark/DCGC-5863/2020\|EPI_ISL_621050\|2020 | .................... | .................... | .................... | .................... | .................... | .................... |
| P | 3788 | hCoV-19/Denmark/DCGC-5864/2020\|EPI_ISL_621051\|2020 | .................... | .................... | .................... | .................... | .................... | .................... |
| P | 3789 | hCoV-19/Denmark/DCGC-5865/2020\|EPI_ISL_621052\|2020 | .................... | .................... | .................... | .................... | .................... | .................... |
| P | 3790 | hCoV-19/Denmark/DCGC-5866/2020\|EPI_ISL_621053\|2020 | .................... | .................... | .................... | .................... | .................... | .................... |
| P | 3791 | hCoV-19/Denmark/DCGC-5867/2020\|EPI_ISL_621054\|2020 | .................... | .................... | .................... | .................... | .................... | .................... |
| P | 3792 | hCoV-19/Denmark/DCGC-5869/2020\|EPI_ISL_621056\|2020 | .................... | .................... | .................... | .................... | .................... | .................... |
| P | 3793 | hCoV-19/Denmark/DCGC-5870/2020\|EPI_ISL_621057\|2020 | .................... | .................... | .................... | .................... | .................... | .................... |
| P | 3794 | hCoV-19/Denmark/DCGC-5871/2020\|EPI_ISL_621058\|2020 | .................... | .................... | .................... | .................... | .................... | .................... |
| P | 3795 | hCoV-19/Denmark/DCGC-5872/2020\|EPI_ISL_621059\|2020 | .................... | .................... | .................... | .................... | .................... | .................... |
| P | 3796 | hCoV-19/Denmark/DCGC-5873/2020\|EPI_ISL_621060\|2020 | .................... | .................... | .................... | .................... | .................... | .................... |
| P | 3797 | hCoV-19/Denmark/DCGC-5874/2020\|EPI_ISL_621061\|2020 | .................... | .................... | .................... | .................... | .................... | .................... |

| | | | | | | | | |
|---|---|---|---|---|---|---|---|---|
| P | 3798 | hCoV-19/Denmark/DCGC-5875/2020\|EPI_ISL_621062\|2020 | ···················· | ···················· | ····················· | ···················· | ···················· | ···················· |
| P | 3799 | hCoV-19/Denmark/DCGC-5876/2020\|EPI_ISL_621063\|2020 | ···················· | ···················· | ····················· | ···················· | ···················· | ···················· |
| P | 3800 | hCoV-19/Denmark/DCGC-5877/2020\|EPI_ISL_621064\|2020 | ···················· | ···················· | ····················· | ···················· | ···················· | ···················· |
| P | 3801 | hCoV-19/Denmark/DCGC-5878/2020\|EPI_ISL_621065\|2020 | ···················· | ···················· | ····················· | ···················· | ···················· | ···················· |
| P | 3802 | hCoV-19/Denmark/DCGC-5879/2020\|EPI_ISL_621066\|2020 | ···················· | ···················· | ····················· | ···················· | ···················· | ···················· |
| P | 3803 | hCoV-19/Denmark/DCGC-5880/2020\|EPI_ISL_621067\|2020 | ···················· | ···················· | ····················· | ···················· | ···················· | ···················· |
| P | 3804 | hCoV-19/Denmark/DCGC-5881/2020\|EPI_ISL_621068\|2020 | ···················· | ···················· | ····················· | ···················· | ···················· | ···················· |
| P | 3805 | hCoV-19/Denmark/DCGC-5882/2020\|EPI_ISL_621069\|2020 | ···················· | ···················· | ····················· | ···················· | ···················· | ···················· |
| P | 3806 | hCoV-19/Denmark/DCGC-5961/2020\|EPI_ISL_621070\|2020 | ···················· | ···················· | ····················· | ···················· | ···················· | ···················· |
| P | 3807 | hCoV-19/Denmark/DCGC-5962/2020\|EPI_ISL_621071\|2020 | ···················· | ···················· | ····················· | ···················· | ···················· | ···················· |
| P | 3808 | hCoV-19/Denmark/DCGC-5963/2020\|EPI_ISL_621072\|2020 | ···················· | ···················· | ····················· | ···················· | ···················· | ···················· |
| P | 3809 | hCoV-19/Denmark/DCGC-5964/2020\|EPI_ISL_621073\|2020 | ···················· | ···················· | ····················· | ···················· | ···················· | ···················· |
| P | 3810 | hCoV-19/Denmark/DCGC-5965/2020\|EPI_ISL_621074\|2020 | ···················· | ···················· | ····················· | ···················· | ···················· | ···················· |
| P | 3811 | hCoV-19/Denmark/DCGC-5966/2020\|EPI_ISL_621075\|2020 | ···················· | ···················· | ····················· | ···················· | ···················· | ···················· |
| P | 3812 | hCoV-19/Denmark/DCGC-5967/2020\|EPI_ISL_621076\|2020 | ···················· | ···················· | ····················· | ···················· | ···················· | ···················· |
| P | 3813 | hCoV-19/Denmark/DCGC-5968/2020\|EPI_ISL_621077\|2020 | ···················· | ···················· | ····················· | ···················· | ···················· | ···················· |
| P | 3814 | hCoV-19/Denmark/DCGC-5969/2020\|EPI_ISL_621078\|2020 | ···················· | ···················· | ····················· | ···················· | ···················· | ···················· |
| P | 3815 | hCoV-19/Denmark/DCGC-5970/2020\|EPI_ISL_621079\|2020 | ···················· | ···················· | ····················· | ···················· | ···················· | ···················· |
| P | 3816 | hCoV-19/Denmark/DCGC-5971/2020\|EPI_ISL_621080\|2020 | ···················· | ···················· | ····················· | ···················· | ···················· | ···················· |
| P | 3817 | hCoV-19/Denmark/DCGC-5972/2020\|EPI_ISL_621081\|2020 | ···················· | ···················· | ····················· | ···················· | ···················· | ···················· |
| P | 3818 | hCoV-19/Denmark/DCGC-5973/2020\|EPI_ISL_621082\|2020 | ···················· | ···················· | ····················· | ···················· | ···················· | ···················· |
| P | 3819 | hCoV-19/Denmark/DCGC-5974/2020\|EPI_ISL_621083\|2020 | ···················· | ···················· | ····················· | ···················· | ···················· | ···················· |
| P | 3820 | hCoV-19/Denmark/DCGC-5977/2020\|EPI_ISL_621086\|2020 | ···················· | ···················· | ····················· | ···················· | ···················· | ···················· |
| P | 3821 | hCoV-19/Denmark/DCGC-5978/2020\|EPI_ISL_621087\|2020 | ···················· | ···················· | ····················· | ···················· | ···················· | ···················· |
| P | 3822 | hCoV-19/Denmark/DCGC-5979/2020\|EPI_ISL_621088\|2020 | ···················· | ···················· | ····················· | ···················· | ···················· | ···················· |
| P | 3823 | hCoV-19/Denmark/DCGC-5980/2020\|EPI_ISL_621089\|2020 | ···················· | ···················· | ····················· | ···················· | ···················· | ···················· |
| P | 3824 | hCoV-19/Denmark/DCGC-6703/2020\|EPI_ISL_622380\|2020 | ···················· | ···················· | ····················· | ···················· | ···················· | ···················· |
| P | 3825 | hCoV-19/Denmark/DCGC-6714/2020\|EPI_ISL_622381\|2020 | ···················· | ···················· | ····················· | ···················· | ···················· | ···················· |
| P | 3826 | hCoV-19/Denmark/DCGC-6733/2020\|EPI_ISL_622383\|2020 | ···················· | ···················· | ····················· | ···················· | ···················· | ···················· |
| P | 3827 | hCoV-19/Denmark/DCGC-6739/2020\|EPI_ISL_622384\|2020 | ···················· | ···················· | ····················· | ···················· | ···················· | ···················· |
| P | 3828 | hCoV-19/Denmark/DCGC-6743/2020\|EPI_ISL_622385\|2020 | ···················· | ···················· | ····················· | ···················· | ···················· | ···················· |
| P | 3829 | hCoV-19/Denmark/DCGC-6704/2020\|EPI_ISL_622386\|2020 | ···················· | ···················· | ····················· | ···················· | ···················· | ···················· |
| P | 3830 | hCoV-19/Denmark/DCGC-6709/2020\|EPI_ISL_622387\|2020 | ···················· | ···················· | ····················· | ···················· | ···················· | ···················· |
| P | 3831 | hCoV-19/Denmark/DCGC-6715/2020\|EPI_ISL_622388\|2020 | ···················· | ···················· | ····················· | ···················· | ···················· | ···················· |
| P | 3832 | hCoV-19/Denmark/DCGC-6720/2020\|EPI_ISL_622389\|2020 | ···················· | ···················· | ····················· | ···················· | ···················· | ···················· |
| P | 3833 | hCoV-19/Denmark/DCGC-6727/2020\|EPI_ISL_622390\|2020 | ···················· | ···················· | ····················· | ···················· | ···················· | ···················· |
| P | 3834 | hCoV-19/Denmark/DCGC-6740/2020\|EPI_ISL_622391\|2020 | ···················· | ···················· | ····················· | ···················· | ···················· | ···················· |
| P | 3835 | hCoV-19/Denmark/DCGC-6744/2020\|EPI_ISL_622392\|2020 | ···················· | ···················· | ····················· | ···················· | ···················· | ···················· |
| P | 3836 | hCoV-19/Denmark/DCGC-6752/2020\|EPI_ISL_622393\|2020 | ···················· | ···················· | ····················· | ···················· | ···················· | ···················· |
| P | 3837 | hCoV-19/Denmark/DCGC-6755/2020\|EPI_ISL_622394\|2020 | ···················· | ···················· | ····················· | ···················· | ···················· | ···················· |
| P | 3838 | hCoV-19/Denmark/DCGC-6710/2020\|EPI_ISL_622395\|2020 | ···················· | ···················· | ····················· | ···················· | ···················· | ···················· |
| P | 3839 | hCoV-19/Denmark/DCGC-6716/2020\|EPI_ISL_622396\|2020 | ···················· | ···················· | ····················· | ···················· | ···················· | ···················· |
| P | 3840 | hCoV-19/Denmark/DCGC-6728/2020\|EPI_ISL_622398\|2020 | ···················· | ···················· | ····················· | ···················· | ···················· | ···················· |
| P | 3841 | hCoV-19/Denmark/DCGC-5239/2020\|EPI_ISL_615170\|2020 | ···················· | ···················· | ····················· | ···················· | ···················· | ···················· |
| P | 3842 | hCoV-19/Denmark/DCGC-5240/2020\|EPI_ISL_615171\|2020 | ···················· | ···················· | ····················· | ···················· | ···················· | ···················· |
| P | 3843 | hCoV-19/Denmark/DCGC-5241/2020\|EPI_ISL_615172\|2020 | ···················· | ···················· | ····················· | ···················· | ···················· | ···················· |
| P | 3844 | hCoV-19/Denmark/DCGC-5242/2020\|EPI_ISL_615173\|2020 | ···················· | ···················· | ····················· | ···················· | ···················· | ···················· |
| P | 3845 | hCoV-19/Denmark/DCGC-5243/2020\|EPI_ISL_615174\|2020 | ···················· | ···················· | ····················· | ···················· | ···················· | ···················· |
| P | 3846 | hCoV-19/Denmark/DCGC-5244/2020\|EPI_ISL_615175\|2020 | ···················· | ···················· | ····················· | ···················· | ···················· | ···················· |
| P | 3847 | hCoV-19/Denmark/DCGC-5245/2020\|EPI_ISL_615176\|2020 | ···················· | ···················· | ····················· | ···················· | ···················· | ···················· |
| P | 3848 | hCoV-19/Denmark/DCGC-5246/2020\|EPI_ISL_615177\|2020 | ···················· | ···················· | ····················· | ···················· | ···················· | ···················· |
| P | 3849 | hCoV-19/Denmark/DCGC-5247/2020\|EPI_ISL_615178\|2020 | ···················· | ···················· | ····················· | ···················· | ···················· | ···················· |
| P | 3850 | hCoV-19/Denmark/DCGC-5248/2020\|EPI_ISL_615179\|2020 | ···················· | ···················· | ····················· | ···················· | ···················· | ···················· |
| P | 3851 | hCoV-19/Denmark/DCGC-5249/2020\|EPI_ISL_615180\|2020 | ···················· | ···················· | ····················· | ···················· | ···················· | ···················· |
| P | 3852 | hCoV-19/Denmark/DCGC-5250/2020\|EPI_ISL_615181\|2020 | ···················· | ···················· | ····················· | ···················· | ···················· | ···················· |
| P | 3853 | hCoV-19/Denmark/DCGC-5251/2020\|EPI_ISL_615182\|2020 | ···················· | ···················· | ····················· | ···················· | ···················· | ···················· |
| P | 3854 | hCoV-19/Denmark/DCGC-5252/2020\|EPI_ISL_615183\|2020 | ···················· | ···················· | ····················· | ···················· | ···················· | ···················· |
| P | 3855 | hCoV-19/Denmark/DCGC-5253/2020\|EPI_ISL_615184\|2020 | ···················· | ···················· | ····················· | ···················· | ···················· | ···················· |
| P | 3856 | hCoV-19/Denmark/DCGC-5254/2020\|EPI_ISL_615185\|2020 | ···················· | ···················· | ····················· | ···················· | ···················· | ···················· |
| P | 3857 | hCoV-19/Denmark/DCGC-5255/2020\|EPI_ISL_615186\|2020 | ···················· | ···················· | ····················· | ···················· | ···················· | ···················· |
| P | 3858 | hCoV-19/Denmark/DCGC-5256/2020\|EPI_ISL_615187\|2020 | ···················· | ···················· | ····················· | ···················· | ···················· | ···················· |
| P | 3859 | hCoV-19/Denmark/DCGC-5257/2020\|EPI_ISL_615188\|2020 | ···················· | ···················· | ····················· | ···················· | ···················· | ···················· |
| P | 3860 | hCoV-19/Denmark/DCGC-5258/2020\|EPI_ISL_615189\|2020 | ···················· | ···················· | ····················· | ···················· | ···················· | ···················· |
| P | 3861 | hCoV-19/Denmark/DCGC-5259/2020\|EPI_ISL_615190\|2020 | ···················· | ···················· | ····················· | ···················· | ···················· | ···················· |
| P | 3862 | hCoV-19/Denmark/DCGC-5260/2020\|EPI_ISL_615191\|2020 | ···················· | ···················· | ····················· | ···················· | ···················· | ···················· |
| P | 3863 | hCoV-19/Denmark/DCGC-5261/2020\|EPI_ISL_615192\|2020 | ···················· | ···················· | ····················· | ···················· | ···················· | ···················· |
| P | 3864 | hCoV-19/Denmark/DCGC-5262/2020\|EPI_ISL_615193\|2020 | ···················· | ···················· | ····················· | ···················· | ···················· | ···················· |
| P | 3865 | hCoV-19/Denmark/DCGC-5263/2020\|EPI_ISL_615194\|2020 | ···················· | ···················· | ····················· | ···················· | ···················· | ···················· |
| P | 3866 | hCoV-19/Denmark/DCGC-5264/2020\|EPI_ISL_615195\|2020 | ···················· | ···················· | ····················· | ···················· | ···················· | ···················· |
| P | 3867 | hCoV-19/Denmark/DCGC-5265/2020\|EPI_ISL_615196\|2020 | ···················· | ···················· | ····················· | ···················· | ···················· | ···················· |
| P | 3868 | hCoV-19/Denmark/DCGC-5266/2020\|EPI_ISL_615197\|2020 | ···················· | ···················· | ····················· | ···················· | ···················· | ···················· |
| P | 3869 | hCoV-19/Denmark/DCGC-5267/2020\|EPI_ISL_615198\|2020 | ···················· | ···················· | ····················· | ···················· | ···················· | ···················· |
| P | 3870 | hCoV-19/Denmark/DCGC-5268/2020\|EPI_ISL_615199\|2020 | ···················· | ···················· | ····················· | ···················· | ···················· | ···················· |
| P | 3871 | hCoV-19/Denmark/DCGC-5300/2020\|EPI_ISL_615231\|2020 | ········T··········· | ···················· | ····················· | ···················· | ···················· | ···················· |
| P | 3872 | hCoV-19/Denmark/DCGC-5310/2020\|EPI_ISL_615241\|2020 | ···················· | ···················· | ····················· | ···················· | ···················· | ···················· |
| P | 3873 | hCoV-19/Denmark/DCGC-5311/2020\|EPI_ISL_615242\|2020 | ···················· | ···················· | ····················· | ···················· | ···················· | ···················· |
| P | 3874 | hCoV-19/Denmark/DCGC-5312/2020\|EPI_ISL_615243\|2020 | ···················· | ···················· | ····················· | ···················· | ···················· | ···················· |
| P | 3875 | hCoV-19/Denmark/DCGC-5313/2020\|EPI_ISL_615244\|2020 | ···················· | ···················· | ····················· | ···················· | ···················· | ···················· |
| P | 3876 | hCoV-19/Denmark/DCGC-5315/2020\|EPI_ISL_615246\|2020 | ···················· | ···················· | ····················· | ···················· | ···················· | ···················· |
| P | 3877 | hCoV-19/Denmark/DCGC-5316/2020\|EPI_ISL_615247\|2020 | ···················· | ···················· | ····················· | ···················· | ···················· | ···················· |

| | | | | | | | | |
|---|---|---|---|---|---|---|---|---|
| P | 3878 | hCoV-19/Denmark/DCGC-5317/2020\|EPI_ISL_615248\|2020 | ···················· | ···················· | ····················· | ···················· | ···················· | ···················· |
| P | 3879 | hCoV-19/Denmark/DCGC-5318/2020\|EPI_ISL_615249\|2020 | ···················· | ···················· | ····················· | ···················· | ···················· | ···················· |
| P | 3880 | hCoV-19/Denmark/DCGC-2868/2020\|EPI_ISL_616562\|2020 | ···················· | ·A·················· | ····················· | ···················· | ···················· | ···················· |
| P | 3881 | hCoV-19/Denmark/DCGC-2875/2020\|EPI_ISL_616570\|2020 | ···················· | ·A·················· | ····················· | ···················· | ···················· | ···················· |
| P | 3882 | hCoV-19/Denmark/DCGC-2876/2020\|EPI_ISL_616571\|2020 | ···················· | ···················· | ····················· | ···················· | ···················· | ···················· |
| P | 3883 | hCoV-19/Denmark/DCGC-2877/2020\|EPI_ISL_616572\|2020 | ···················· | ···················· | ····················· | ···················· | ···················· | ···················· |
| P | 3884 | hCoV-19/Denmark/DCGC-2878/2020\|EPI_ISL_616573\|2020 | ···················· | ···················· | ····················· | ···················· | ···················· | ···················· |
| P | 3885 | hCoV-19/Denmark/DCGC-2879/2020\|EPI_ISL_616574\|2020 | ···················· | ···················· | ····················· | ···················· | ···················· | ···················· |
| P | 3886 | hCoV-19/Denmark/DCGC-2880/2020\|EPI_ISL_616575\|2020 | ···················· | ·A·················· | ····················· | ···················· | ···················· | ···················· |
| P | 3887 | hCoV-19/Denmark/DCGC-2881/2020\|EPI_ISL_616576\|2020 | ···················· | ·A·················· | ····················· | ···················· | ···················· | ···················· |
| P | 3888 | hCoV-19/Denmark/DCGC-2882/2020\|EPI_ISL_616577\|2020 | ···················· | ·A·················· | ····················· | ···················· | ···················· | ··A················· |
| P | 3889 | hCoV-19/Denmark/DCGC-2883/2020\|EPI_ISL_616578\|2020 | ···················· | ···················· | ····················· | ···················· | ···················· | ···················· |
| P | 3890 | hCoV-19/Denmark/DCGC-2884/2020\|EPI_ISL_616579\|2020 | ···················· | ···················· | ····················· | ···················· | ···················· | ···················· |
| P | 3891 | hCoV-19/Denmark/DCGC-2672/2020\|EPI_ISL_617892\|2020 | ···················· | ···················· | ····················· | ···················· | ···················· | ···················· |
| P | 3892 | hCoV-19/Denmark/DCGC-5822/2020\|EPI_ISL_621009\|2020 | ···················· | ···················· | ····················· | ···················· | ···················· | ···················· |
| P | 3893 | hCoV-19/Denmark/DCGC-5823/2020\|EPI_ISL_621010\|2020 | ···················· | ···················· | ····················· | ···················· | ···················· | ···················· |
| P | 3894 | hCoV-19/Denmark/DCGC-5825/2020\|EPI_ISL_621012\|2020 | ···················· | ···················· | ····················· | ···················· | ···················· | ···················· |
| P | 3895 | hCoV-19/Denmark/DCGC-5826/2020\|EPI_ISL_621013\|2020 | ···················· | ···················· | ····················· | ···················· | ···················· | ···················· |
| P | 3896 | hCoV-19/Denmark/DCGC-5827/2020\|EPI_ISL_621014\|2020 | ···················· | ···················· | ····················· | ···················· | ···················· | ···················· |
| P | 3897 | hCoV-19/Denmark/DCGC-5828/2020\|EPI_ISL_621015\|2020 | ···················· | ···················· | ····················· | ···················· | ···················· | ···················· |
| P | 3898 | hCoV-19/Denmark/DCGC-5829/2020\|EPI_ISL_621016\|2020 | ···················· | ···················· | ····················· | ···················· | ···················· | ···················· |
| P | 3899 | hCoV-19/Denmark/DCGC-5831/2020\|EPI_ISL_621018\|2020 | ···················· | ···················· | ····················· | ···················· | ···················· | ···················· |
| P | 3900 | hCoV-19/Denmark/DCGC-5832/2020\|EPI_ISL_621019\|2020 | ···················· | ···················· | ····················· | ···················· | ···················· | ···················· |
| P | 3901 | hCoV-19/Denmark/DCGC-5834/2020\|EPI_ISL_621021\|2020 | ···················· | ···················· | ····················· | ···················· | ···················· | ···················· |
| P | 3902 | hCoV-19/Denmark/DCGC-5835/2020\|EPI_ISL_621022\|2020 | ···················· | ···················· | ····················· | ···················· | ···················· | ···················· |
| P | 3903 | hCoV-19/Denmark/DCGC-5836/2020\|EPI_ISL_621023\|2020 | ···················· | ···················· | ····················· | ···················· | ···················· | ···················· |
| P | 3904 | hCoV-19/Denmark/DCGC-5837/2020\|EPI_ISL_621024\|2020 | ···················· | ···················· | ····················· | ···················· | ···················· | ···················· |
| P | 3905 | hCoV-19/Denmark/DCGC-5838/2020\|EPI_ISL_621025\|2020 | ···················· | ···················· | ····················· | ···················· | ···················· | ···················· |
| P | 3906 | hCoV-19/Denmark/DCGC-5839/2020\|EPI_ISL_621026\|2020 | ···················· | ···················· | ····················· | ···················· | ···················· | ···················· |
| P | 3907 | hCoV-19/Denmark/DCGC-5840/2020\|EPI_ISL_621027\|2020 | ···················· | ···················· | ····················· | ···················· | ···················· | ···················· |
| P | 3908 | hCoV-19/Denmark/DCGC-5842/2020\|EPI_ISL_621029\|2020 | ···················· | ···················· | ····················· | ···················· | ···················· | ···················· |
| P | 3909 | hCoV-19/Denmark/DCGC-5843/2020\|EPI_ISL_621030\|2020 | ···················· | ···················· | ····················· | ···················· | ···················· | ···················· |
| P | 3910 | hCoV-19/Denmark/DCGC-5844/2020\|EPI_ISL_621031\|2020 | ···················· | ···················· | ····················· | ···················· | ···················· | ···················· |
| P | 3911 | hCoV-19/Denmark/DCGC-5845/2020\|EPI_ISL_621032\|2020 | ···················· | ···················· | ····················· | ···················· | ···················· | ···················· |
| P | 3912 | hCoV-19/Denmark/DCGC-5846/2020\|EPI_ISL_621033\|2020 | ···················· | ···················· | ····················· | ···················· | ···················· | ···················· |
| P | 3913 | hCoV-19/Denmark/DCGC-5847/2020\|EPI_ISL_621034\|2020 | ···················· | ···················· | ····················· | ···················· | ···················· | ···················· |
| P | 3914 | hCoV-19/Denmark/DCGC-5848/2020\|EPI_ISL_621035\|2020 | ···················· | ···················· | ····················· | ···················· | ···················· | ···················· |
| P | 3915 | hCoV-19/Denmark/DCGC-5849/2020\|EPI_ISL_621036\|2020 | ···················· | ···················· | ····················· | ···················· | ···················· | ···················· |
| P | 3916 | hCoV-19/Denmark/DCGC-5850/2020\|EPI_ISL_621037\|2020 | ···················· | ···················· | ····················· | ···················· | ···················· | ···················· |
| P | 3917 | hCoV-19/Denmark/DCGC-5851/2020\|EPI_ISL_621038\|2020 | ···················· | ···················· | ····················· | ···················· | ···················· | ···················· |
| P | 3918 | hCoV-19/Denmark/DCGC-5853/2020\|EPI_ISL_621040\|2020 | ···················· | ···················· | ····················· | ···················· | ···················· | ···················· |
| P | 3919 | hCoV-19/Denmark/DCGC-5854/2020\|EPI_ISL_621041\|2020 | ···················· | ···················· | ····················· | ···················· | ···················· | ···················· |
| P | 3920 | hCoV-19/Denmark/DCGC-5855/2020\|EPI_ISL_621042\|2020 | ···················· | ···················· | ····················· | ···················· | ···················· | ···················· |
| P | 3921 | hCoV-19/Denmark/DCGC-5856/2020\|EPI_ISL_621043\|2020 | ···················· | ···················· | ····················· | ···················· | ···················· | ···················· |
| P | 3922 | hCoV-19/Denmark/DCGC-5857/2020\|EPI_ISL_621044\|2020 | ···················· | ···················· | ····················· | ···················· | ···················· | ···················· |
| P | 3923 | hCoV-19/Denmark/DCGC-5858/2020\|EPI_ISL_621045\|2020 | ···················· | ···················· | ····················· | ···················· | ···················· | ···················· |
| P | 3924 | hCoV-19/Denmark/DCGC-5859/2020\|EPI_ISL_621046\|2020 | ···················· | ···················· | ····················· | ···················· | ···················· | ···················· |
| P | 3925 | hCoV-19/Denmark/DCGC-5860/2020\|EPI_ISL_621047\|2020 | ···················· | ···················· | ····················· | ···················· | ···················· | ···················· |
| P | 3926 | hCoV-19/Denmark/DCGC-5862/2020\|EPI_ISL_621049\|2020 | ···················· | ···················· | ····················· | ···················· | ···················· | ···················· |
| P | 3927 | hCoV-19/Denmark/DCGC-6358/2020\|EPI_ISL_622305\|2020 | ···················· | ···················· | ····················· | ···················· | ···················· | ···················· |
| P | 3928 | hCoV-19/Denmark/DCGC-6370/2020\|EPI_ISL_622306\|2020 | ···················· | ···················· | ····················· | ···················· | ···················· | ···················· |
| P | 3929 | hCoV-19/Denmark/DCGC-6376/2020\|EPI_ISL_622307\|2020 | ···················· | ···················· | ····················· | ···················· | ···················· | ···················· |
| P | 3930 | hCoV-19/Denmark/DCGC-6383/2020\|EPI_ISL_622308\|2020 | ···················· | ···················· | ····················· | ···················· | ···················· | ···················· |
| P | 3931 | hCoV-19/Denmark/DCGC-6341/2020\|EPI_ISL_622309\|2020 | ···················· | ···················· | ····················· | ···················· | ···················· | ···················· |
| P | 3932 | hCoV-19/Denmark/DCGC-6348/2020\|EPI_ISL_622310\|2020 | ···················· | ···················· | ····················· | ···················· | ···················· | ···················· |
| P | 3933 | hCoV-19/Denmark/DCGC-6359/2020\|EPI_ISL_622311\|2020 | ···················· | ···················· | ····················· | ···················· | ···················· | ···················· |
| P | 3934 | hCoV-19/Denmark/DCGC-6371/2020\|EPI_ISL_622312\|2020 | ···················· | ···················· | ····················· | ···················· | ···················· | ···················· |
| P | 3935 | hCoV-19/Denmark/DCGC-6377/2020\|EPI_ISL_622313\|2020 | ···················· | ···················· | ····················· | ···················· | ···················· | ···················· |
| P | 3936 | hCoV-19/Denmark/DCGC-6384/2020\|EPI_ISL_622314\|2020 | ···················· | ···················· | ····················· | ···················· | ···················· | ···················· |
| P | 3937 | hCoV-19/Denmark/DCGC-6392/2020\|EPI_ISL_622315\|2020 | ···················· | ···················· | ····················· | ···················· | ···················· | ···················· |
| P | 3938 | hCoV-19/Denmark/DCGC-6374/2020\|EPI_ISL_622338\|2020 | ···················· | ···················· | ····················· | ···················· | ···················· | ···················· |
| P | 3939 | hCoV-19/Denmark/DCGC-6388/2020\|EPI_ISL_622339\|2020 | ···················· | ···················· | ····················· | ···················· | ···················· | ···················· |
| P | 3940 | hCoV-19/Denmark/DCGC-6395/2020\|EPI_ISL_622340\|2020 | ···················· | ···················· | ····················· | ···················· | ···················· | ···················· |
| P | 3941 | hCoV-19/Denmark/DCGC-6399/2020\|EPI_ISL_622341\|2020 | ···················· | ···················· | ····················· | ···················· | ···················· | ···················· |
| P | 3942 | hCoV-19/Denmark/DCGC-6717/2020\|EPI_ISL_622343\|2020 | ···················· | ···················· | ····················· | ···················· | ···················· | ···················· |
| P | 3943 | hCoV-19/Denmark/DCGC-6729/2020\|EPI_ISL_622344\|2020 | ···················· | ···················· | ····················· | ···················· | ···················· | ···················· |
| P | 3944 | hCoV-19/Denmark/DCGC-6734/2020\|EPI_ISL_622345\|2020 | ···················· | ···················· | ····················· | ···················· | ···················· | ···················· |
| P | 3945 | hCoV-19/Denmark/DCGC-6741/2020\|EPI_ISL_622346\|2020 | ···················· | ···················· | ····················· | ···················· | ···················· | ···················· |
| P | 3946 | hCoV-19/Denmark/DCGC-6749/2020\|EPI_ISL_622347\|2020 | ···················· | ···················· | ····················· | ···················· | ···················· | ···················· |
| P | 3947 | hCoV-19/Denmark/DCGC-6757/2020\|EPI_ISL_622348\|2020 | ···················· | ···················· | ····················· | ···················· | ···················· | ···················· |
| P | 3948 | hCoV-19/Denmark/DCGC-6700/2020\|EPI_ISL_622349\|2020 | ···················· | ···················· | ····················· | ···················· | ···················· | ···················· |
| P | 3949 | hCoV-19/Denmark/DCGC-6706/2020\|EPI_ISL_622350\|2020 | ···················· | ···················· | ····················· | ···················· | ···················· | ···················· |
| P | 3950 | hCoV-19/Denmark/DCGC-6711/2020\|EPI_ISL_622351\|2020 | ···················· | ···················· | ····················· | ···················· | ···················· | ···················· |
| P | 3951 | hCoV-19/Denmark/DCGC-6718/2020\|EPI_ISL_622352\|2020 | ········T··········· | ···················· | ···········T········· | ···················· | ···················· | ···················· |
| P | 3952 | hCoV-19/Denmark/DCGC-6722/2020\|EPI_ISL_622353\|2020 | ···················· | ···················· | ····················· | ···················· | ···················· | ···················· |
| P | 3953 | hCoV-19/Denmark/DCGC-6730/2020\|EPI_ISL_622354\|2020 | ···················· | ···················· | ····················· | ···················· | ···················· | ···················· |
| P | 3954 | hCoV-19/Denmark/DCGC-6742/2020\|EPI_ISL_622356\|2020 | ···················· | ···················· | ····················· | ···················· | ···················· | ···················· |
| P | 3955 | hCoV-19/Denmark/DCGC-6746/2020\|EPI_ISL_622357\|2020 | ···················· | ···················· | ····················· | ···················· | ···················· | ···················· |
| P | 3956 | hCoV-19/Denmark/DCGC-6750/2020\|EPI_ISL_622358\|2020 | ···················· | ···················· | ····················· | ···················· | ···················· | ···················· |
| P | 3957 | hCoV-19/Denmark/DCGC-6712/2020\|EPI_ISL_622360\|2020 | ···················· | ···················· | ····················· | ···················· | ···················· | ···················· |

| | | | | | | | | |
|---|---|---|---|---|---|---|---|---|
| P | 3958 | hCoV-19/Denmark/DCGC-6723/2020\|EPI_ISL_622361\|2020 | ···················· | ···················· | ···················· | ···················· | ···················· | ···················· |
| P | 3959 | hCoV-19/Denmark/DCGC-6736/2020\|EPI_ISL_622362\|2020 | ···················· | ···················· | ···················· | ···················· | ···················· | ···················· |
| P | 3960 | hCoV-19/Denmark/DCGC-6701/2020\|EPI_ISL_622364\|2020 | ···················· | ···················· | ···················· | ···················· | ···················· | ···················· |
| P | 3961 | hCoV-19/Denmark/DCGC-6731/2020\|EPI_ISL_622367\|2020 | ···················· | ···················· | ···················· | ···················· | ···················· | ···················· |
| P | 3962 | hCoV-19/Denmark/DCGC-6737/2020\|EPI_ISL_622368\|2020 | ···················· | ···················· | ···················· | ···················· | ···················· | ···················· |
| P | 3963 | hCoV-19/Denmark/DCGC-6759/2020\|EPI_ISL_622369\|2020 | ···················· | ···················· | ···················· | ···················· | ···················· | ···················· |
| P | 3964 | hCoV-19/Denmark/DCGC-6702/2020\|EPI_ISL_622370\|2020 | ···················· | ···················· | ···················· | ···················· | ···················· | ···················· |
| P | 3965 | hCoV-19/Denmark/DCGC-6708/2020\|EPI_ISL_622371\|2020 | ···················· | ···················· | ···················· | ···················· | ···················· | ···················· |
| P | 3966 | hCoV-19/Denmark/DCGC-6725/2020\|EPI_ISL_622373\|2020 | ···················· | ···················· | ···················· | ···················· | ···················· | ···················· |
| P | 3967 | hCoV-19/Denmark/DCGC-6732/2020\|EPI_ISL_622374\|2020 | ···················· | ···················· | ···················· | ···················· | ···················· | ···················· |
| P | 3968 | hCoV-19/Denmark/DCGC-6747/2020\|EPI_ISL_622376\|2020 | ···················· | ···················· | ···················· | ···················· | ···················· | ···················· |
| P | 3969 | hCoV-19/Denmark/DCGC-6751/2020\|EPI_ISL_622377\|2020 | ···················· | ···················· | ···················· | ···················· | ···················· | ···················· |
| P | 3970 | hCoV-19/Denmark/DCGC-6754/2020\|EPI_ISL_622378\|2020 | ···················· | ···················· | ···················· | ···················· | ···················· | ···················· |
| P | 3971 | hCoV-19/Denmark/DCGC-6760/2020\|EPI_ISL_622379\|2020 | ···················· | ···················· | ···················· | ···················· | ···················· | ···················· |
| P | 3972 | hCoV-19/England/CAMC-AB117A/2020\|EPI_ISL_623600\|20 | ···················· | ···················· | ···················· | ···················· | ···················· | ···················· |
| P | 3973 | hCoV-19/England/MILK-AB88F6/2020\|EPI_ISL_623601\|20 | ···················· | ···················· | ···················· | ···················· | ···················· | ···················· |
| P | 3974 | hCoV-19/England/MILK-AB8814/2020\|EPI_ISL_623603\|20 | ···················· | ···················· | ···················· | ···················· | ·····T·············· | ···················· |
| P | 3975 | hCoV-19/England/MILK-AB796D/2020\|EPI_ISL_623604\|20 | ···················· | ···················· | ···················· | ···················· | ···················· | ···················· |
| P | 3976 | hCoV-19/England/CAMC-AB1DA3/2020\|EPI_ISL_623605\|20 | ···················· | ···················· | ···················· | ···················· | ···················· | ···················· |
| P | 3977 | hCoV-19/England/MILK-AB7E13/2020\|EPI_ISL_623606\|20 | ···················· | ···················· | ···················· | ···················· | ···················· | ···················· |
| P | 3978 | hCoV-19/England/MILK-AB7E04/2020\|EPI_ISL_623607\|20 | ···················· | ···················· | ···················· | ···················· | ·····T·············· | ···············A···· |
| P | 3979 | hCoV-19/England/CAMC-AB21F1/2020\|EPI_ISL_623608\|20 | ···················· | ···················· | ···················· | ···················· | ···················· | ···················· |
| P | 3980 | hCoV-19/England/CAMC-AB1D49/2020\|EPI_ISL_623609\|20 | ···················· | ···················· | ···················· | ···················· | ·····T·············· | ···················· |
| P | 3981 | hCoV-19/England/MILK-AB7930/2020\|EPI_ISL_623610\|20 | ···················· | ···················· | ···················· | ···················· | ···················· | ···················· |
| P | 3982 | hCoV-19/England/CAMC-AB1857/2020\|EPI_ISL_623611\|20 | ···················· | ···················· | ···················· | ···················· | ···················· | ···················· |
| P | 3983 | hCoV-19/England/CAMC-AB1E64/2020\|EPI_ISL_623612\|20 | ···················· | ···················· | ···················· | ···················· | ···················· | ···················· |
| P | 3984 | hCoV-19/England/MILK-AB8638/2020\|EPI_ISL_623614\|20 | ···················· | ···················· | ···················· | ···················· | ···················· | ···················· |
| P | 3985 | hCoV-19/England/MILK-AB85A4/2020\|EPI_ISL_623615\|20 | ···················· | ···················· | ···················· | ···················· | ···················· | ···················· |
| P | 3986 | hCoV-19/England/CAMC-AB262F/2020\|EPI_ISL_623616\|20 | ···················· | ···················· | ···················· | ···················· | ···················· | ···················· |
| P | 3987 | hCoV-19/England/CAMC-AB1DD0/2020\|EPI_ISL_623617\|20 | ···················· | ···················· | ···················· | ···················· | ···················· | ···················· |
| P | 3988 | hCoV-19/England/CAMC-AB1532/2020\|EPI_ISL_623619\|20 | ···················· | ···················· | ···················· | ···················· | ···················· | ···················· |
| P | 3989 | hCoV-19/England/MILK-AB8911/2020\|EPI_ISL_623620\|20 | ···················· | ···················· | ···················· | ···················· | ···················· | ···················· |
| P | 3990 | hCoV-19/England/MILK-AB7CEC/2020\|EPI_ISL_623621\|20 | ···················· | ···················· | ···················· | ···················· | ···················· | ···················· |
| P | 3991 | hCoV-19/England/MILK-AB8780/2020\|EPI_ISL_623622\|20 | ···················· | ···················· | ···················· | ···················· | ···················· | ···················· |
| P | 3992 | hCoV-19/England/MILK-AB7912/2020\|EPI_ISL_623623\|20 | ···················· | ···················· | ···················· | ···················· | ···················· | ···················· |
| P | 3993 | hCoV-19/England/MILK-AB8629/2020\|EPI_ISL_623625\|20 | ···················· | ···················· | ···················· | ···················· | ·····T·············· | ···················· |
| P | 3994 | hCoV-19/England/CAMC-AB1893/2020\|EPI_ISL_623626\|20 | ···················· | ···················· | ···················· | ···················· | ···················· | ···················· |
| P | 3995 | hCoV-19/England/MILK-AB8DE8/2020\|EPI_ISL_623627\|20 | ···················· | ···················· | ···················· | ···················· | ···················· | ···················· |
| P | 3996 | hCoV-19/England/MILK-AB7BC1/2020\|EPI_ISL_623628\|20 | ···················· | ···················· | ···················· | ···················· | ···················· | ···················· |
| P | 3997 | hCoV-19/England/MILK-AB7C19/2020\|EPI_ISL_623629\|20 | ···················· | ···················· | ···················· | ···················· | ···················· | ···················· |
| P | 3998 | hCoV-19/England/MILK-AB753C/2020\|EPI_ISL_623636\|20 | ···················· | ···················· | ···················· | ···················· | ···················· | ···················· |
| P | 3999 | hCoV-19/England/MILK-AB7B94/2020\|EPI_ISL_623637\|20 | ···················· | ·A·················· | ···················· | ···················· | ···················· | ···A················ |
| P | 4000 | hCoV-19/England/MILK-AB8805/2020\|EPI_ISL_623638\|20 | ···················· | ·A·················· | ···················· | ···················· | ···················· | ···················· |
| P | 4001 | hCoV-19/England/CAMC-AB114D/2020\|EPI_ISL_623640\|20 | ···················· | ···················· | ···················· | ···················· | ···················· | ···················· |
| P | 4002 | hCoV-19/England/CAMC-AB1918/2020\|EPI_ISL_623641\|20 | ···················· | ···················· | ···················· | ···················· | ···················· | ···················· |
| P | 4003 | hCoV-19/England/MILK-AB9075/2020\|EPI_ISL_623642\|20 | ···················· | ···················· | ···················· | ···················· | ···················· | ···················· |
| P | 4004 | hCoV-19/England/CAMC-AB1CB5/2020\|EPI_ISL_623643\|20 | ···················· | ···················· | ···················· | ···················· | ···················· | ···················· |
| P | 4005 | hCoV-19/England/CAMC-AB113E/2020\|EPI_ISL_623644\|20 | ···················· | ···················· | ···················· | ···················· | ···················· | ···················· |
| P | 4006 | hCoV-19/England/CAMC-AB1523/2020\|EPI_ISL_623645\|20 | ···················· | ···················· | ···················· | ···················· | ···················· | ···················· |
| P | 4007 | hCoV-19/England/MILK-AB8DCA/2020\|EPI_ISL_623646\|20 | ···················· | ···················· | ···················· | ···················· | ···················· | ···················· |
| P | 4008 | hCoV-19/England/MILK-AB7D16/2020\|EPI_ISL_623669\|20 | ···················· | ···················· | ···················· | ···················· | ···················· | ···················· |
| P | 4009 | hCoV-19/England/MILK-AB87DB/2020\|EPI_ISL_623670\|20 | ···················· | ···················· | ···················· | ···················· | ···················· | ···················· |
| P | 4010 | hCoV-19/England/MILK-AB85D1/2020\|EPI_ISL_623671\|20 | ···················· | ···················· | ···················· | ···················· | ···················· | ···················· |
| P | 4011 | hCoV-19/England/MILK-AB88C9/2020\|EPI_ISL_623672\|20 | ···················· | ···················· | ···················· | ···················· | ·····T·············· | ···················· |
| P | 4012 | hCoV-19/England/MILK-AB937C/2020\|EPI_ISL_623673\|20 | ···················· | ···················· | ···················· | ···················· | ···················· | ···················· |
| P | 4013 | hCoV-19/England/CAMC-AB1365/2020\|EPI_ISL_623675\|20 | ···················· | ···················· | ···················· | ···················· | ···················· | ···················· |
| P | 4014 | hCoV-19/England/MILK-AB8E4F/2020\|EPI_ISL_623676\|20 | ···················· | ···················· | ···················· | ···················· | ···················· | ···················· |
| P | 4015 | hCoV-19/England/CAMC-AB1D85/2020\|EPI_ISL_623679\|20 | ···················· | ·A·················· | ···················· | ···················· | ···················· | ···A················ |
| P | 4016 | hCoV-19/England/CAMC-AB1CD3/2020\|EPI_ISL_623680\|20 | ···················· | ···················· | ···················· | ···················· | ···················· | ···················· |
| P | 4017 | hCoV-19/England/MILK-AB7D61/2020\|EPI_ISL_623681\|20 | ···················· | ···················· | ···················· | ···················· | ···················· | ···················· |
| P | 4018 | hCoV-19/England/MILK-AB9093/2020\|EPI_ISL_623682\|20 | ···················· | ···················· | ···················· | ···················· | ···················· | ···················· |
| P | 4019 | hCoV-19/England/CAMC-AB18C0/2020\|EPI_ISL_623683\|20 | ···················· | ···················· | ···················· | ···················· | ···················· | ···················· |
| P | 4020 | hCoV-19/England/MILK-AB901B/2020\|EPI_ISL_623684\|20 | ···················· | ···················· | ···················· | ···················· | ···················· | ···················· |
| P | 4021 | hCoV-19/England/MILK-AB9B92/2020\|EPI_ISL_623686\|20 | ···················· | ···················· | ···················· | ···················· | ···················· | ···················· |
| P | 4022 | hCoV-19/England/MILK-AB879F/2020\|EPI_ISL_623687\|20 | ···················· | ···················· | ···················· | ···················· | ···················· | ···················· |
| P | 4023 | hCoV-19/England/CAMC-AB1514/2020\|EPI_ISL_623688\|20 | ···················· | ···················· | ···················· | ···················· | ···················· | ···················· |
| P | 4024 | hCoV-19/England/MILK-AB7B58/2020\|EPI_ISL_623689\|20 | ···················· | ···················· | ···················· | ···················· | ···················· | ···················· |
| P | 4025 | hCoV-19/England/CAMC-AB12A4/2020\|EPI_ISL_623690\|20 | ···················· | ···················· | ···················· | ···················· | ···················· | ···················· |
| P | 4026 | hCoV-19/England/MILK-AB8920/2020\|EPI_ISL_623691\|20 | ···················· | ···················· | ···················· | ···················· | ·····T·············· | ···················· |
| P | 4027 | hCoV-19/England/MILK-AB7A1F/2020\|EPI_ISL_623692\|20 | ···················· | ···················· | ···················· | ···················· | ···················· | ···················· |
| P | 4028 | hCoV-19/England/MILK-AB8D24/2020\|EPI_ISL_623693\|20 | ···················· | ···················· | ···················· | ···················· | ···················· | ···················· |
| P | 4029 | hCoV-19/England/CAMC-AB1C1F/2020\|EPI_ISL_623694\|20 | ···················· | ···················· | ···················· | ···················· | ···················· | ···················· |
| P | 4030 | hCoV-19/England/MILK-AB7CA0/2020\|EPI_ISL_623695\|20 | ···················· | ···················· | ···················· | ···················· | ···················· | ···················· |
| P | 4031 | hCoV-19/England/MILK-AB9B74/2020\|EPI_ISL_623697\|20 | ···················· | ···················· | ···················· | ···················· | ···················· | ···················· |
| P | 4032 | hCoV-19/England/MILK-AB79D6/2020\|EPI_ISL_623698\|20 | ···················· | ···················· | ···················· | ···················· | ·····T·············· | ···················· |
| P | 4033 | hCoV-19/England/MILK-AB9B65/2020\|EPI_ISL_623699\|20 | ···················· | ···················· | ···················· | ···················· | ···················· | ···················· |
| P | 4034 | hCoV-19/England/ALDP-A952C4/2020\|EPI_ISL_624900\|20 | ···················· | ···················· | ···················· | ···················· | ···················· | ···················· |
| P | 4035 | hCoV-19/England/ALDP-A9475D/2020\|EPI_ISL_624901\|20 | ···················· | ···················· | ···················· | ···················· | ···················· | ···················· |
| P | 4036 | hCoV-19/England/ALDP-A96436/2020\|EPI_ISL_624902\|20 | ···················· | ···················· | ···················· | ···················· | ···················· | ···················· |
| P | 4037 | hCoV-19/England/ALDP-A94535/2020\|EPI_ISL_624904\|20 | ···················· | ···················· | ···················· | ···················· | ···················· | ···················· |

| | | | | | | | | |
|---|---|---|---|---|---|---|---|---|
| P | 4038 | hCoV-19/England/ALDP-A962D2/2020\|EPI_ISL_624905\|20 | ···················· | ···················· | ···················· | ···················· | ···················· | ···················· |
| P | 4039 | hCoV-19/England/ALDP-A94E76/2020\|EPI_ISL_624906\|20 | ···················· | ···················· | ···················· | ···················· | ···················· | ···················· |
| P | 4040 | hCoV-19/England/ALDP-AA0EA4/2020\|EPI_ISL_624908\|20 | ···················· | ···················· | ···················· | ···················· | ···················· | ···················· |
| P | 4041 | hCoV-19/England/ALDP-AA3287/2020\|EPI_ISL_624910\|20 | ···················· | ···················· | ···················· | ···················· | ···················· | ···················· |
| P | 4042 | hCoV-19/England/ALDP-AA2288/2020\|EPI_ISL_624912\|20 | ···················· | ···················· | ···················· | ···················· | ···················· | ···················· |
| P | 4043 | hCoV-19/Wales/ALDP-AA4981/2020\|EPI_ISL_624913\|2020 | ···················· | ···················· | ···················· | ···················· | ···················· | ···················· |
| P | 4044 | hCoV-19/England/ALDP-AA2130/2020\|EPI_ISL_624914\|20 | ···················· | ·A·················· | ···················· | ···················· | ···················· | ··A················· |
| P | 4045 | hCoV-19/England/ALDP-AA0A37/2020\|EPI_ISL_624915\|20 | ···················· | ···················· | ···················· | ···················· | ···················· | ···················· |
| P | 4046 | hCoV-19/England/ALDP-AA3867/2020\|EPI_ISL_624916\|20 | ···················· | ···················· | ···················· | ···················· | ···················· | ···················· |
| P | 4047 | hCoV-19/England/ALDP-AA182D/2020\|EPI_ISL_624917\|20 | ···················· | ···················· | ···················· | ···················· | ···················· | ···················· |
| P | 4048 | hCoV-19/England/ALDP-AA562F/2020\|EPI_ISL_624918\|20 | ···················· | ·A·················· | ···················· | ···················· | ···················· | ··A················· |
| P | 4049 | hCoV-19/England/ALDP-AA3269/2020\|EPI_ISL_624919\|20 | ···················· | ···················· | ···················· | ···················· | ···················· | ···················· |
| P | 4050 | hCoV-19/England/ALDP-AA35BB/2020\|EPI_ISL_624921\|20 | ···················· | ···················· | ···················· | ···················· | ···················· | ···················· |
| P | 4051 | hCoV-19/England/ALDP-AA3F17/2020\|EPI_ISL_624922\|20 | ···················· | ···················· | ···················· | ···················· | ···················· | ···················· |
| P | 4052 | hCoV-19/England/ALDP-AA5294/2020\|EPI_ISL_624923\|20 | ···················· | ···················· | ···················· | ···················· | ···················· | ···················· |
| P | 4053 | hCoV-19/England/ALDP-AA17E4/2020\|EPI_ISL_624924\|20 | ···················· | ···················· | ···················· | ···················· | ···················· | ···················· |
| P | 4054 | hCoV-19/England/ALDP-AA49FA/2020\|EPI_ISL_624925\|20 | ···················· | ···················· | ···················· | ···················· | ···················· | ···················· |
| P | 4055 | hCoV-19/England/ALDP-AA168D/2020\|EPI_ISL_624926\|20 | ···················· | ···················· | ···················· | ···················· | ···················· | ···················· |
| P | 4056 | hCoV-19/England/ALDP-AA0B34/2020\|EPI_ISL_624927\|20 | ···················· | ···················· | ···················· | ···················· | ···················· | ···················· |
| P | 4057 | hCoV-19/England/ALDP-AA0E95/2020\|EPI_ISL_624928\|20 | ···················· | ···················· | ···················· | ···················· | ···················· | ···················· |
| P | 4058 | hCoV-19/England/ALDP-AA3894/2020\|EPI_ISL_624929\|20 | ···················· | ···················· | ···················· | ···················· | ···················· | ···················· |
| P | 4059 | hCoV-19/England/ALDP-AA44EA/2020\|EPI_ISL_624930\|20 | ···················· | ···················· | ···················· | ···················· | ···················· | ···················· |
| P | 4060 | hCoV-19/England/ALDP-AA574A/2020\|EPI_ISL_624931\|20 | ···················· | ···················· | ···················· | ···················· | ···················· | ···················· |
| P | 4061 | hCoV-19/England/ALDP-AA33DF/2020\|EPI_ISL_624932\|20 | ···················· | ···················· | ···················· | ···················· | ···················· | ···················· |
| P | 4062 | hCoV-19/England/ALDP-AA2BD8/2020\|EPI_ISL_624933\|20 | ···················· | ···················· | ···················· | ···················· | ···················· | ···················· |
| P | 4063 | hCoV-19/England/ALDP-AA0897/2020\|EPI_ISL_624934\|20 | ···················· | ···················· | ···················· | ···················· | ···················· | ···················· |
| P | 4064 | hCoV-19/England/ALDP-AA4541/2020\|EPI_ISL_624935\|20 | ···················· | ···················· | ···················· | ···················· | ···················· | ···················· |
| P | 4065 | hCoV-19/England/ALDP-AA3603/2020\|EPI_ISL_624936\|20 | ···················· | ···················· | ···················· | ···················· | ···················· | ···················· |
| P | 4066 | hCoV-19/England/ALDP-AA3876/2020\|EPI_ISL_624937\|20 | ···················· | ···················· | ···················· | ···················· | ···················· | ···················· |
| P | 4067 | hCoV-19/England/ALDP-AA55AA/2020\|EPI_ISL_624938\|20 | ···················· | ···················· | ···················· | ···················· | ···················· | ···················· |
| P | 4068 | hCoV-19/England/ALDP-AA52C1/2020\|EPI_ISL_624939\|20 | ···················· | ···················· | ···················· | ···················· | ···················· | ···················· |
| P | 4069 | hCoV-19/England/ALDP-AA3621/2020\|EPI_ISL_624940\|20 | ···················· | ·A·················· | ···················· | ···················· | ···················· | ··A················· |
| P | 4070 | hCoV-19/England/ALDP-AA0D4D/2020\|EPI_ISL_624942\|20 | ···················· | ···················· | ···················· | ···················· | ···················· | ···················· |
| P | 4071 | hCoV-19/England/ALDP-AA0B25/2020\|EPI_ISL_624943\|20 | ···················· | ···················· | ···················· | ···················· | ···················· | ···················· |
| P | 4072 | hCoV-19/England/ALDP-AA3F53/2020\|EPI_ISL_624944\|20 | ···················· | ···················· | ···················· | ···················· | ···················· | ···················· |
| P | 4073 | hCoV-19/England/ALDP-AA322D/2020\|EPI_ISL_624945\|20 | ···················· | ···················· | ···················· | ···················· | ···················· | ···················· |
| P | 4074 | hCoV-19/England/ALDP-AA1E67/2020\|EPI_ISL_624946\|20 | ···················· | ···················· | ···················· | ···················· | ···················· | ···················· |
| P | 4075 | hCoV-19/England/ALDP-AA53CE/2020\|EPI_ISL_624947\|20 | ···················· | ···················· | ···················· | ···················· | ···················· | ···················· |
| P | 4076 | hCoV-19/England/ALDP-AA0B07/2020\|EPI_ISL_624949\|20 | ···················· | ···················· | ···················· | ···················· | ···················· | ···················· |
| P | 4077 | hCoV-19/England/ALDP-AA2BC9/2020\|EPI_ISL_624950\|20 | ···················· | ···················· | ···················· | ···················· | ···················· | ··A················· |
| P | 4078 | hCoV-19/England/ALDP-AA17B7/2020\|EPI_ISL_624951\|20 | ···················· | ···················· | ···················· | ···················· | ···················· | ···················· |
| P | 4079 | hCoV-19/England/ALDP-AA174E/2020\|EPI_ISL_624952\|20 | ···················· | ···················· | ···················· | ···················· | ···················· | ···················· |
| P | 4080 | hCoV-19/England/ALDP-AA08B5/2020\|EPI_ISL_624953\|20 | ···················· | ·A·················· | ···················· | ···················· | ···················· | ···················· |
| P | 4081 | hCoV-19/England/ALDP-AA20BB/2020\|EPI_ISL_624955\|20 | ···················· | ···················· | ···················· | ···················· | ···················· | ···················· |
| P | 4082 | hCoV-19/England/ALDP-AA0B9E/2020\|EPI_ISL_624956\|20 | ···················· | ···················· | ···················· | ···················· | ···················· | ···················· |
| P | 4083 | hCoV-19/England/ALDP-AA2877/2020\|EPI_ISL_624957\|20 | ···················· | ···················· | ···················· | ···················· | ···················· | ···················· |
| P | 4084 | hCoV-19/England/ALDP-AA37B5/2020\|EPI_ISL_624958\|20 | ···················· | ···················· | ···················· | ···················· | ···················· | ···················· |
| P | 4085 | hCoV-19/England/ALDP-AA33B1/2020\|EPI_ISL_624959\|20 | ···················· | ···················· | ···················· | ···················· | ···················· | ···················· |
| P | 4086 | hCoV-19/England/ALDP-AA0C22/2020\|EPI_ISL_624967\|20 | ···················· | ···················· | ···················· | ···················· | ···················· | ···················· |
| P | 4087 | hCoV-19/England/ALDP-AA1DB5/2020\|EPI_ISL_624968\|20 | ···················· | ···················· | ···················· | ···················· | ···················· | ···················· |
| P | 4088 | hCoV-19/England/ALDP-AA37A6/2020\|EPI_ISL_624969\|20 | ···················· | ···················· | ···················· | ···················· | ···················· | ···················· |
| P | 4089 | hCoV-19/England/ALDP-AA3612/2020\|EPI_ISL_624970\|20 | ···················· | ·A·················· | ···················· | ···················· | ···················· | ··A················· |
| P | 4090 | hCoV-19/England/ALDP-AA2C02/2020\|EPI_ISL_624971\|20 | ···················· | ···················· | ···················· | ···················· | ···················· | ···················· |
| P | 4091 | hCoV-19/England/ALDP-AA5416/2020\|EPI_ISL_624972\|20 | ···················· | ···················· | ···················· | ···················· | ···················· | ···················· |
| P | 4092 | hCoV-19/England/ALDP-AA185A/2020\|EPI_ISL_624973\|20 | ···················· | ···················· | ···················· | ···················· | ···················· | ···················· |
| P | 4093 | hCoV-19/England/ALDP-AA20D9/2020\|EPI_ISL_624974\|20 | ···················· | ···················· | ···················· | ···················· | ···················· | ···················· |
| P | 4094 | hCoV-19/England/ALDP-AA1140/2020\|EPI_ISL_624975\|20 | ···················· | ···················· | ···················· | ···················· | ···················· | ···················· |
| P | 4095 | hCoV-19/England/ALDP-AA1614/2020\|EPI_ISL_624976\|20 | ···················· | ···················· | ···················· | ···················· | ···················· | ···················· |
| P | 4096 | hCoV-19/England/ALDP-AA0C8C/2020\|EPI_ISL_624977\|20 | ···················· | ···················· | ···················· | ···················· | ···················· | ···················· |
| P | 4097 | hCoV-19/USA/TX-DSHS-1101/2020\|EPI_ISL_615139\|2020- | ···················· | ···················· | ···················· | ···················· | ···················· | ···················· |
| P | 4098 | hCoV-19/USA/TX-DSHS-1121/2020\|EPI_ISL_615158\|2020- | ···················· | ·A·················· | ···················· | ···················· | ···················· | ···················· |
| P | 4099 | hCoV-19/Denmark/DCGC-5235/2020\|EPI_ISL_615166\|2020 | ···················· | ···················· | ···················· | ···················· | ···················· | ···················· |
| P | 4100 | hCoV-19/Denmark/DCGC-5236/2020\|EPI_ISL_615167\|2020 | ···················· | ···················· | ···················· | ···················· | ···················· | ···················· |
| P | 4101 | hCoV-19/Denmark/DCGC-5237/2020\|EPI_ISL_615168\|2020 | ···················· | ···················· | ···················· | ···················· | ···················· | ···················· |
| P | 4102 | hCoV-19/Denmark/DCGC-5238/2020\|EPI_ISL_615169\|2020 | ···················· | ···················· | ···················· | ···················· | ···················· | ···················· |
| P | 4103 | hCoV-19/Denmark/DCGC-6302/2020\|EPI_ISL_616400\|2020 | ···················· | ···················· | ···················· | ···················· | ···················· | ···················· |
| P | 4104 | hCoV-19/Denmark/DCGC-3630/2020\|EPI_ISL_616401\|2020 | ···················· | ···················· | ···················· | ···················· | ···················· | ···················· |
| P | 4105 | hCoV-19/Denmark/DCGC-3632/2020\|EPI_ISL_616403\|2020 | ···················· | ···················· | ···················· | ···················· | ···················· | ···················· |
| P | 4106 | hCoV-19/Denmark/DCGC-3633/2020\|EPI_ISL_616404\|2020 | ···················· | ···················· | ···················· | ···················· | ···················· | ···················· |
| P | 4107 | hCoV-19/Denmark/DCGC-3634/2020\|EPI_ISL_616405\|2020 | ···················· | ···················· | ···················· | ···················· | ···················· | ···················· |
| P | 4108 | hCoV-19/Denmark/DCGC-2575/2020\|EPI_ISL_616406\|2020 | ···················· | ···················· | ···················· | ···················· | ···················· | ···················· |
| P | 4109 | hCoV-19/Denmark/DCGC-2576/2020\|EPI_ISL_616407\|2020 | ···················· | ···················· | ···················· | ···················· | ···················· | ···················· |
| P | 4110 | hCoV-19/Denmark/DCGC-2578/2020\|EPI_ISL_616409\|2020 | ···················· | ···················· | ···················· | ···················· | ···················· | ···················· |
| P | 4111 | hCoV-19/Denmark/DCGC-2894/2020\|EPI_ISL_616410\|2020 | ···················· | ·A·················· | ···················· | ···················· | ···················· | ···················· |
| P | 4112 | hCoV-19/Denmark/DCGC-2895/2020\|EPI_ISL_616411\|2020 | ···················· | ···················· | ···················· | ···················· | ···················· | ···················· |
| P | 4113 | hCoV-19/Denmark/DCGC-2896/2020\|EPI_ISL_616412\|2020 | ···················· | ···················· | ···················· | ···················· | ···················· | ···················· |
| P | 4114 | hCoV-19/Denmark/DCGC-2897/2020\|EPI_ISL_616413\|2020 | ···················· | ···················· | ···················· | ···················· | ···················· | ···················· |
| P | 4115 | hCoV-19/Denmark/DCGC-2898/2020\|EPI_ISL_616414\|2020 | ···················· | ···················· | ···················· | ···················· | ···················· | ···················· |
| P | 4116 | hCoV-19/Denmark/DCGC-2579/2020\|EPI_ISL_616415\|2020 | ···················· | ·········A·········· | ···················· | ···················· | ···················· | ···················· |
| P | 4117 | hCoV-19/Denmark/DCGC-1713/2020\|EPI_ISL_616416\|2020 | ···················· | ···················· | ···················· | ···················· | ···················· | ···················· |

| | | | | | | | | |
|---|---|---|---|---|---|---|---|---|
| P | 4118 | hCoV-19/Denmark/DCGC-2580/2020\|EPI_ISL_616417\|2020 | ···················· | ···················· | ····················· | ···················· | ···················· | ···················· |
| P | 4119 | hCoV-19/Denmark/DCGC-2581/2020\|EPI_ISL_616418\|2020 | ···················· | ···················· | ····················· | ···················· | ···················· | ···················· |
| P | 4120 | hCoV-19/Denmark/DCGC-2582/2020\|EPI_ISL_616419\|2020 | ···················· | ···················· | ····················· | ···················· | ···················· | ···················· |
| P | 4121 | hCoV-19/Denmark/DCGC-2583/2020\|EPI_ISL_616420\|2020 | ···················· | ···················· | ····················· | ···················· | ···················· | ···················· |
| P | 4122 | hCoV-19/Denmark/DCGC-2584/2020\|EPI_ISL_616421\|2020 | ···················· | ···················· | ····················· | ···················· | ······T············· | ···················· |
| P | 4123 | hCoV-19/Denmark/DCGC-2585/2020\|EPI_ISL_616422\|2020 | ···················· | ···················· | ····················· | ···················· | ···················· | ···················· |
| P | 4124 | hCoV-19/Denmark/DCGC-2586/2020\|EPI_ISL_616423\|2020 | ···················· | ···················· | ····················· | ···················· | ···················· | ···················· |
| P | 4125 | hCoV-19/Denmark/DCGC-2692/2020\|EPI_ISL_616424\|2020 | ···················· | ···················· | ····················· | ···················· | ···················· | ···················· |
| P | 4126 | hCoV-19/Denmark/DCGC-2693/2020\|EPI_ISL_616425\|2020 | ···················· | ···················· | ····················· | ···················· | ···················· | ···················· |
| P | 4127 | hCoV-19/Denmark/DCGC-2694/2020\|EPI_ISL_616426\|2020 | ···················· | ···················· | ····················· | ···················· | ···················· | ···················· |
| P | 4128 | hCoV-19/Denmark/DCGC-2696/2020\|EPI_ISL_616428\|2020 | ···················· | ···················· | ····················· | ···················· | ···················· | ···················· |
| P | 4129 | hCoV-19/Denmark/DCGC-2697/2020\|EPI_ISL_616429\|2020 | ···················· | ···················· | ····················· | ···················· | ···················· | ···················· |
| P | 4130 | hCoV-19/Denmark/DCGC-2824/2020\|EPI_ISL_616460\|2020 | ···················· | ···················· | ····················· | ···················· | ···················· | ···················· |
| P | 4131 | hCoV-19/Denmark/DCGC-2825/2020\|EPI_ISL_616461\|2020 | ···················· | ···················· | ····················· | ···················· | ······T············· | ···················· |
| P | 4132 | hCoV-19/Denmark/DCGC-2573/2020\|EPI_ISL_616462\|2020 | ···················· | ···················· | ····················· | ···················· | ···················· | ···················· |
| P | 4133 | hCoV-19/Denmark/DCGC-2718/2020\|EPI_ISL_616463\|2020 | ···················· | ···················· | ···········T········· | ···················· | ···················· | ···················· |
| P | 4134 | hCoV-19/Denmark/DCGC-2719/2020\|EPI_ISL_616464\|2020 | ···················· | ···················· | ····················· | ···················· | ···················· | ···················· |
| P | 4135 | hCoV-19/Denmark/DCGC-2720/2020\|EPI_ISL_616465\|2020 | ···················· | ···················· | ····················· | ···················· | ···················· | ···················· |
| P | 4136 | hCoV-19/Denmark/DCGC-2721/2020\|EPI_ISL_616466\|2020 | ···················· | ·A··A··············· | ····················· | ···················· | ···················· | ···················· |
| P | 4137 | hCoV-19/Denmark/DCGC-2722/2020\|EPI_ISL_616467\|2020 | ···················· | ·A··A··············· | ····················· | ···················· | ···················· | ···················· |
| P | 4138 | hCoV-19/Denmark/DCGC-2723/2020\|EPI_ISL_616468\|2020 | ···················· | ···················· | ····················· | ···················· | ···················· | ···············A···· |
| P | 4139 | hCoV-19/Denmark/DCGC-2725/2020\|EPI_ISL_616470\|2020 | ···················· | ···················· | ····················· | ···················· | ···················· | ···················· |
| P | 4140 | hCoV-19/Denmark/DCGC-2726/2020\|EPI_ISL_616471\|2020 | ···················· | ···················· | ····················· | ···················· | ···················· | ···················· |
| P | 4141 | hCoV-19/Denmark/DCGC-2727/2020\|EPI_ISL_616472\|2020 | ···················· | ···················· | ····················· | ···················· | ···················· | ···················· |
| P | 4142 | hCoV-19/Denmark/DCGC-2728/2020\|EPI_ISL_616473\|2020 | ···················· | ···················· | ····················· | ···················· | ···················· | ···················· |
| P | 4143 | hCoV-19/Denmark/DCGC-2729/2020\|EPI_ISL_616474\|2020 | ···················· | ···················· | ····················· | ···················· | ···················· | ···················· |
| P | 4144 | hCoV-19/Denmark/DCGC-2730/2020\|EPI_ISL_616475\|2020 | ···················· | ···················· | ····················· | ···················· | ···················· | ···················· |
| P | 4145 | hCoV-19/Denmark/DCGC-2732/2020\|EPI_ISL_616477\|2020 | ···················· | ···················· | ····················· | ···················· | ······T············· | ···················· |
| P | 4146 | hCoV-19/Denmark/DCGC-2733/2020\|EPI_ISL_616478\|2020 | ···················· | ·A·················· | ····················· | ···················· | ···················· | ···················· |
| P | 4147 | hCoV-19/Denmark/DCGC-2734/2020\|EPI_ISL_616479\|2020 | ···················· | ······C············· | ····················· | ···················· | ···················· | ···················· |
| P | 4148 | hCoV-19/Denmark/DCGC-2736/2020\|EPI_ISL_616481\|2020 | ········T··········· | ···················· | ····················· | ···················· | ···················· | ···················· |
| P | 4149 | hCoV-19/Denmark/DCGC-2737/2020\|EPI_ISL_616482\|2020 | ···················· | ·A·················· | ····················· | ···················· | ···················· | ···················· |
| P | 4150 | hCoV-19/Denmark/DCGC-2738/2020\|EPI_ISL_616483\|2020 | ···················· | ···················· | ····················· | ···················· | ···················· | ···················· |
| P | 4151 | hCoV-19/Denmark/DCGC-2739/2020\|EPI_ISL_616484\|2020 | ···················· | ···················· | ····················· | ···················· | ···················· | ···················· |
| P | 4152 | hCoV-19/Denmark/DCGC-2740/2020\|EPI_ISL_616485\|2020 | ···················· | ···················· | ····················· | ···················· | ···················· | ···················· |
| P | 4153 | hCoV-19/Denmark/DCGC-2741/2020\|EPI_ISL_616486\|2020 | ···················· | ·A··A··············· | ····················· | ···················· | ···················· | ···················· |
| P | 4154 | hCoV-19/Denmark/DCGC-2742/2020\|EPI_ISL_616487\|2020 | ···················· | ···················· | ····················· | ···················· | ···················· | ···················· |
| P | 4155 | hCoV-19/Denmark/DCGC-2743/2020\|EPI_ISL_616488\|2020 | ···················· | ···················· | ····················· | ···················· | ···················· | ···················· |
| P | 4156 | hCoV-19/Denmark/DCGC-2744/2020\|EPI_ISL_616489\|2020 | ···················· | ···················· | ····················· | ···················· | ···················· | ···················· |
| P | 4157 | hCoV-19/Denmark/DCGC-2745/2020\|EPI_ISL_616490\|2020 | ···················· | ···················· | ····················· | ···················· | ···················· | ···················· |
| P | 4158 | hCoV-19/Denmark/DCGC-2746/2020\|EPI_ISL_616491\|2020 | ···················· | ···················· | ····················· | ···················· | ···················· | ···················· |
| P | 4159 | hCoV-19/Denmark/DCGC-2747/2020\|EPI_ISL_616492\|2020 | ···················· | ···················· | ····················· | ···················· | ···················· | ···················· |
| P | 4160 | hCoV-19/Denmark/DCGC-2748/2020\|EPI_ISL_616493\|2020 | ···················· | ···················· | ····················· | ···················· | ···················· | ···················· |
| P | 4161 | hCoV-19/Denmark/DCGC-2749/2020\|EPI_ISL_616494\|2020 | ···················· | ·A·················· | ····················· | ···················· | ···················· | ···················· |
| P | 4162 | hCoV-19/Denmark/DCGC-2750/2020\|EPI_ISL_616495\|2020 | ···················· | ···················· | ····················· | ···················· | ···················· | ···················· |
| P | 4163 | hCoV-19/Denmark/DCGC-2751/2020\|EPI_ISL_616496\|2020 | ···················· | ···················· | ····················· | ···················· | ···················· | ···················· |
| P | 4164 | hCoV-19/Denmark/DCGC-2752/2020\|EPI_ISL_616497\|2020 | ···················· | ···················· | ····················· | ···················· | ···················· | ···················· |
| P | 4165 | hCoV-19/Denmark/DCGC-2754/2020\|EPI_ISL_616499\|2020 | ···················· | ···················· | ····················· | ···················· | ···················· | ···················· |
| P | 4166 | hCoV-19/Denmark/DCGC-2360/2020\|EPI_ISL_617700\|2020 | ···················· | ···················· | ····················· | ···················· | ···················· | ···················· |
| P | 4167 | hCoV-19/Denmark/DCGC-2361/2020\|EPI_ISL_617701\|2020 | ···················· | ···················· | ····················· | ···················· | ···················· | ···················· |
| P | 4168 | hCoV-19/Denmark/DCGC-2362/2020\|EPI_ISL_617702\|2020 | ···················· | ···················· | ····················· | ···················· | ···················· | ···················· |
| P | 4169 | hCoV-19/Denmark/DCGC-2363/2020\|EPI_ISL_617703\|2020 | ···················· | ···················· | ····················· | ···················· | ···················· | ···················· |
| P | 4170 | hCoV-19/Denmark/DCGC-2364/2020\|EPI_ISL_617704\|2020 | ···················· | ···················· | ····················· | ···················· | ···················· | ···················· |
| P | 4171 | hCoV-19/Denmark/DCGC-2365/2020\|EPI_ISL_617705\|2020 | ···················· | ···················· | ····················· | ···················· | ···················· | ···················· |
| P | 4172 | hCoV-19/Denmark/DCGC-2366/2020\|EPI_ISL_617706\|2020 | ···················· | ···················· | ····················· | ···················· | ···················· | ···················· |
| P | 4173 | hCoV-19/Denmark/DCGC-2367/2020\|EPI_ISL_617707\|2020 | ···················· | ···················· | ····················· | ···················· | ···················· | ···················· |
| P | 4174 | hCoV-19/Denmark/DCGC-2368/2020\|EPI_ISL_617708\|2020 | ···················· | ···················· | ····················· | ···················· | ···················· | ···················· |
| P | 4175 | hCoV-19/Denmark/DCGC-2369/2020\|EPI_ISL_617709\|2020 | ···················· | ···················· | ····················· | ···················· | ······T············· | ···················· |
| P | 4176 | hCoV-19/Denmark/DCGC-2370/2020\|EPI_ISL_617710\|2020 | ···················· | ···················· | ····················· | ···················· | ···················· | ···················· |
| P | 4177 | hCoV-19/Denmark/DCGC-2371/2020\|EPI_ISL_617711\|2020 | ···················· | ···················· | ····················· | ···················· | ···················· | ···················· |
| P | 4178 | hCoV-19/Denmark/DCGC-2372/2020\|EPI_ISL_617712\|2020 | ···················· | ···················· | ····················· | ···················· | ···················· | ···················· |
| P | 4179 | hCoV-19/Denmark/DCGC-2373/2020\|EPI_ISL_617713\|2020 | ···················· | ···················· | ····················· | ···················· | ···················· | ···················· |
| P | 4180 | hCoV-19/Denmark/DCGC-2374/2020\|EPI_ISL_617714\|2020 | ···················· | ·A·················· | ····················· | ···················· | ···················· | ···················· |
| P | 4181 | hCoV-19/Denmark/DCGC-2899/2020\|EPI_ISL_617715\|2020 | ···················· | ···················· | ····················· | ···················· | ···················· | ···················· |
| P | 4182 | hCoV-19/Denmark/DCGC-2292/2020\|EPI_ISL_617716\|2020 | ···················· | ···················· | ····················· | ···················· | ···················· | ···················· |
| P | 4183 | hCoV-19/Denmark/DCGC-2293/2020\|EPI_ISL_617717\|2020 | ···················· | ·A·················· | ····················· | ···················· | ···················· | ···A················ |
| P | 4184 | hCoV-19/Denmark/DCGC-2294/2020\|EPI_ISL_617718\|2020 | ···················· | ···················· | ····················· | ···················· | ···················· | ···················· |
| P | 4185 | hCoV-19/Denmark/DCGC-2295/2020\|EPI_ISL_617719\|2020 | ···················· | ·A·················· | ····················· | ···················· | ···················· | ···················· |
| P | 4186 | hCoV-19/Denmark/DCGC-2296/2020\|EPI_ISL_617720\|2020 | ···················· | ···················· | ····················· | ···················· | ···················· | ···················· |
| P | 4187 | hCoV-19/Denmark/DCGC-2298/2020\|EPI_ISL_617722\|2020 | ···················· | ···················· | ····················· | ···················· | ···················· | ···················· |
| P | 4188 | hCoV-19/Denmark/DCGC-2685/2020\|EPI_ISL_617723\|2020 | ···················· | ···················· | ····················· | ···················· | ···················· | ···················· |
| P | 4189 | hCoV-19/Denmark/DCGC-2299/2020\|EPI_ISL_617724\|2020 | ···················· | ···················· | ····················· | ···················· | ···················· | ···················· |
| P | 4190 | hCoV-19/Denmark/DCGC-2300/2020\|EPI_ISL_617725\|2020 | ···················· | ···················· | ····················· | ···················· | ···················· | ···················· |
| P | 4191 | hCoV-19/Denmark/DCGC-2301/2020\|EPI_ISL_617726\|2020 | ···················· | ···················· | ····················· | ···················· | ···················· | ···················· |
| P | 4192 | hCoV-19/Denmark/DCGC-2302/2020\|EPI_ISL_617727\|2020 | ···················· | ···················· | ····················· | ···················· | ···················· | ···················· |
| P | 4193 | hCoV-19/Denmark/DCGC-2303/2020\|EPI_ISL_617728\|2020 | ···················· | ···················· | ····················· | ···················· | ···················· | ···················· |
| P | 4194 | hCoV-19/Denmark/DCGC-2304/2020\|EPI_ISL_617729\|2020 | ···················· | ···················· | ····················· | ···················· | ···················· | ···················· |
| P | 4195 | hCoV-19/Denmark/DCGC-2305/2020\|EPI_ISL_617730\|2020 | ···················· | ···················· | ····················· | ···················· | ···················· | ···················· |
| P | 4196 | hCoV-19/Denmark/DCGC-2306/2020\|EPI_ISL_617731\|2020 | ···················· | ···················· | ····················· | ···················· | ···················· | ···················· |
| P | 4197 | hCoV-19/Denmark/DCGC-2308/2020\|EPI_ISL_617733\|2020 | ···················· | ···················· | ····················· | ···················· | ···················· | ···················· |

| | | | | | | | |
|---|---|---|---|---|---|---|---|
| P | 4198 hCoV-19/Denmark/DCGC-2309/2020\|EPI_ISL_617734\|2020 | .................... | .................... | ..................... | .................... | .................... | .................... |
| P | 4199 hCoV-19/Denmark/DCGC-2310/2020\|EPI_ISL_617735\|2020 | .................... | .................... | ..................... | .................... | .................... | .................... |
| P | 4200 hCoV-19/Denmark/DCGC-2311/2020\|EPI_ISL_617736\|2020 | .................... | .................... | ..................... | .................... | .................... | .................... |
| P | 4201 hCoV-19/Denmark/DCGC-2312/2020\|EPI_ISL_617737\|2020 | .................... | .................... | ..................... | .................... | .................... | .................... |
| P | 4202 hCoV-19/Denmark/DCGC-2686/2020\|EPI_ISL_617738\|2020 | .................... | .................... | ..................... | .................... | .................... | .................... |
| P | 4203 hCoV-19/Denmark/DCGC-2687/2020\|EPI_ISL_617739\|2020 | .................... | .................... | ..................... | .................... | .................... | .................... |
| P | 4204 hCoV-19/Denmark/DCGC-2688/2020\|EPI_ISL_617740\|2020 | .................... | .................... | ..................... | .................... | .................... | .................... |
| P | 4205 hCoV-19/Denmark/DCGC-2473/2020\|EPI_ISL_617741\|2020 | .................... | .................... | ..................... | .................... | .....T.............. | .................... |
| P | 4206 hCoV-19/Denmark/DCGC-2474/2020\|EPI_ISL_617742\|2020 | .................... | .................... | ..................... | .................... | .................... | .................... |
| P | 4207 hCoV-19/Denmark/DCGC-2475/2020\|EPI_ISL_617743\|2020 | .................... | .................... | ..................... | .................... | .................... | .................... |
| P | 4208 hCoV-19/Denmark/DCGC-2476/2020\|EPI_ISL_617744\|2020 | .................... | .................... | ..................... | .................... | .................... | .................... |
| P | 4209 hCoV-19/Denmark/DCGC-2477/2020\|EPI_ISL_617745\|2020 | .................... | .................... | ..................... | .................... | .....T.............. | .................... |
| P | 4210 hCoV-19/Denmark/DCGC-2478/2020\|EPI_ISL_617746\|2020 | .................... | .................... | ..................... | .................... | .................... | .................... |
| P | 4211 hCoV-19/Denmark/DCGC-2479/2020\|EPI_ISL_617747\|2020 | .................... | .................... | ..................... | .................... | .................... | .................... |
| P | 4212 hCoV-19/Denmark/DCGC-2480/2020\|EPI_ISL_617748\|2020 | .................... | .................... | ..................... | .................... | .................... | .................... |
| P | 4213 hCoV-19/Denmark/DCGC-2481/2020\|EPI_ISL_617749\|2020 | .................... | .................... | ..................... | .................... | .................... | .................... |
| P | 4214 hCoV-19/Denmark/DCGC-2482/2020\|EPI_ISL_617750\|2020 | .................... | .................... | ..................... | .................... | .................... | .................... |
| P | 4215 hCoV-19/Denmark/DCGC-2484/2020\|EPI_ISL_617752\|2020 | .................... | .................... | ..................... | .................... | .................... | .................... |
| P | 4216 hCoV-19/Denmark/DCGC-2485/2020\|EPI_ISL_617753\|2020 | .................... | .................... | ..................... | .................... | .................... | .................... |
| P | 4217 hCoV-19/Denmark/DCGC-2518/2020\|EPI_ISL_617754\|2020 | .................... | .................... | ..................... | .................... | .................... | .................... |
| P | 4218 hCoV-19/Denmark/DCGC-2519/2020\|EPI_ISL_617755\|2020 | .................... | .................... | ..................... | .................... | .................... | .................... |
| P | 4219 hCoV-19/Denmark/DCGC-2486/2020\|EPI_ISL_617756\|2020 | .................... | .A.................. | ..................... | .................... | .................... | ..A................. |
| P | 4220 hCoV-19/Denmark/DCGC-2565/2020\|EPI_ISL_617757\|2020 | .................... | .................... | ..................... | .................... | .................... | .................... |
| P | 4221 hCoV-19/Denmark/DCGC-2487/2020\|EPI_ISL_617758\|2020 | .................... | .................... | ..................... | .................... | .................... | .................... |
| P | 4222 hCoV-19/Denmark/DCGC-2488/2020\|EPI_ISL_617759\|2020 | .................... | .................... | ..................... | .................... | .................... | .................... |
| P | 4223 hCoV-19/Denmark/DCGC-2521/2020\|EPI_ISL_617791\|2020 | .................... | .................... | ..................... | .................... | .................... | .................... |
| P | 4224 hCoV-19/Denmark/DCGC-2522/2020\|EPI_ISL_617792\|2020 | .................... | .................... | ..................... | .................... | .................... | .................... |
| P | 4225 hCoV-19/Denmark/DCGC-2523/2020\|EPI_ISL_617793\|2020 | .................... | .................... | ..................... | .................... | .................... | .................... |
| P | 4226 hCoV-19/Denmark/DCGC-2524/2020\|EPI_ISL_617794\|2020 | .................... | .................... | ..................... | .................... | .................... | .................... |
| P | 4227 hCoV-19/Denmark/DCGC-2525/2020\|EPI_ISL_617795\|2020 | .................... | .A.................. | ..................... | .................... | .................... | .................... |
| P | 4228 hCoV-19/Denmark/DCGC-2527/2020\|EPI_ISL_617797\|2020 | .................... | .................... | ..................... | .................... | .................... | .................... |
| P | 4229 hCoV-19/Denmark/DCGC-2528/2020\|EPI_ISL_617798\|2020 | .................... | .................... | ..................... | .................... | .................... | .................... |
| P | 4230 hCoV-19/Denmark/DCGC-2529/2020\|EPI_ISL_617799\|2020 | .................... | .................... | ..................... | .................... | .................... | .................... |
| P | 4231 hCoV-19/Denmark/DCGC-5814/2020\|EPI_ISL_621001\|2020 | .................... | .................... | ..................... | .................... | .................... | .................... |
| P | 4232 hCoV-19/Denmark/DCGC-5815/2020\|EPI_ISL_621002\|2020 | .................... | .................... | ..................... | .................... | .................... | .................... |
| P | 4233 hCoV-19/Denmark/DCGC-5816/2020\|EPI_ISL_621003\|2020 | .................... | .................... | ..................... | .................... | .................... | .................... |
| P | 4234 hCoV-19/Denmark/DCGC-5817/2020\|EPI_ISL_621004\|2020 | .................... | .................... | ..................... | .................... | .................... | .................... |
| P | 4235 hCoV-19/Denmark/DCGC-5818/2020\|EPI_ISL_621005\|2020 | .................... | .................... | ..................... | .................... | .................... | .................... |
| P | 4236 hCoV-19/Denmark/DCGC-5819/2020\|EPI_ISL_621006\|2020 | .................... | .................... | ..................... | .................... | .................... | .................... |
| P | 4237 hCoV-19/Denmark/DCGC-4591/2020\|EPI_ISL_622260\|2020 | .................... | .................... | ..................... | .................... | .................... | .................... |
| P | 4238 hCoV-19/Denmark/DCGC-1469/2020\|EPI_ISL_622262\|2020 | .................... | .................... | ..................... | .................... | .................... | .................... |
| P | 4239 hCoV-19/Denmark/DCGC-1471/2020\|EPI_ISL_622263\|2020 | .................... | .................... | ..................... | .................... | .................... | .................... |
| P | 4240 hCoV-19/Denmark/DCGC-1472/2020\|EPI_ISL_622264\|2020 | .................... | .................... | ..................... | .................... | .................... | .................... |
| P | 4241 hCoV-19/Denmark/DCGC-1470/2020\|EPI_ISL_622265\|2020 | .................... | .................... | ..................... | .................... | .................... | .................... |
| P | 4242 hCoV-19/Denmark/DCGC-1473/2020\|EPI_ISL_622266\|2020 | .................... | .................... | ..................... | .................... | .................... | .................... |
| P | 4243 hCoV-19/Denmark/DCGC-1474/2020\|EPI_ISL_622267\|2020 | .................... | .................... | ..................... | .................... | .................... | .................... |
| P | 4244 hCoV-19/Denmark/DCGC-1475/2020\|EPI_ISL_622268\|2020 | .................... | .................... | ..................... | .................... | .................... | .................... |
| P | 4245 hCoV-19/Denmark/DCGC-1477/2020\|EPI_ISL_622269\|2020 | .................... | .................... | ..................... | .................... | .................... | .................... |
| P | 4246 hCoV-19/Denmark/DCGC-1476/2020\|EPI_ISL_622270\|2020 | .................... | .................... | ..................... | .................... | .................... | .................... |
| P | 4247 hCoV-19/Denmark/DCGC-1462/2020\|EPI_ISL_622271\|2020 | .................... | .................... | ..................... | .................... | .................... | .................... |
| P | 4248 hCoV-19/Denmark/DCGC-1463/2020\|EPI_ISL_622272\|2020 | .................... | .................... | ..................... | .................... | .................... | .................... |
| P | 4249 hCoV-19/Denmark/DCGC-1464/2020\|EPI_ISL_622273\|2020 | .................... | .................... | ..................... | .................... | .................... | .................... |
| P | 4250 hCoV-19/Denmark/DCGC-1465/2020\|EPI_ISL_622274\|2020 | .................... | .................... | ..................... | .................... | .................... | .................... |
| P | 4251 hCoV-19/Denmark/DCGC-1466/2020\|EPI_ISL_622275\|2020 | .................... | .................... | ..................... | .................... | .................... | .................... |
| P | 4252 hCoV-19/Denmark/DCGC-1467/2020\|EPI_ISL_622276\|2020 | .................... | .................... | ..................... | .................... | .................... | .................... |
| P | 4253 hCoV-19/Denmark/DCGC-1468/2020\|EPI_ISL_622277\|2020 | .................... | .................... | ..................... | .................... | .................... | .................... |
| P | 4254 hCoV-19/Denmark/DCGC-6356/2020\|EPI_ISL_622278\|2020 | .................... | .................... | ..................... | .................... | .................... | .................... |
| P | 4255 hCoV-19/Denmark/DCGC-6362/2020\|EPI_ISL_622279\|2020 | .................... | .................... | ..................... | .................... | .................... | .................... |
| P | 4256 hCoV-19/Denmark/DCGC-6380/2020\|EPI_ISL_622280\|2020 | .................... | .................... | ..................... | .................... | .................... | .................... |
| P | 4257 hCoV-19/Denmark/DCGC-6389/2020\|EPI_ISL_622281\|2020 | .................... | .................... | ..................... | .................... | .................... | .................... |
| P | 4258 hCoV-19/Denmark/DCGC-6396/2020\|EPI_ISL_622282\|2020 | .................... | .................... | ..................... | .................... | .................... | .................... |
| P | 4259 hCoV-19/Denmark/DCGC-6400/2020\|EPI_ISL_622283\|2020 | .................... | .................... | ..................... | .................... | .................... | .................... |
| P | 4260 hCoV-19/Denmark/DCGC-6339/2020\|EPI_ISL_622284\|2020 | .................... | .................... | ..................... | .................... | .................... | .................... |
| P | 4261 hCoV-19/Denmark/DCGC-6345/2020\|EPI_ISL_622285\|2020 | .................... | .................... | ..................... | .................... | .................... | .................... |
| P | 4262 hCoV-19/Denmark/DCGC-6352/2020\|EPI_ISL_622286\|2020 | .................... | .................... | ..................... | .................... | .................... | .................... |
| P | 4263 hCoV-19/Denmark/DCGC-6357/2020\|EPI_ISL_622287\|2020 | .................... | .................... | ..................... | .................... | .................... | .................... |
| P | 4264 hCoV-19/Denmark/DCGC-6363/2020\|EPI_ISL_622288\|2020 | .................... | .................... | ..................... | .................... | .................... | .................... |
| P | 4265 hCoV-19/Denmark/DCGC-6368/2020\|EPI_ISL_622289\|2020 | .................... | .................... | ..................... | .................... | .................... | .................... |
| P | 4266 hCoV-19/Denmark/DCGC-6375/2020\|EPI_ISL_622290\|2020 | .................... | .................... | ..................... | .................... | .................... | .................... |
| P | 4267 hCoV-19/Denmark/DCGC-6381/2020\|EPI_ISL_622291\|2020 | .................... | .................... | ..................... | .................... | .................... | .................... |
| P | 4268 hCoV-19/Denmark/DCGC-6386/2020\|EPI_ISL_622292\|2020 | .................... | .................... | ..................... | .................... | .................... | .................... |
| P | 4269 hCoV-19/Denmark/DCGC-6390/2020\|EPI_ISL_622293\|2020 | .................... | .................... | ..................... | .................... | .................... | .................... |
| P | 4270 hCoV-19/Denmark/DCGC-6397/2020\|EPI_ISL_622294\|2020 | .................... | .................... | ..................... | .................... | .................... | .................... |
| P | 4271 hCoV-19/Denmark/DCGC-6401/2020\|EPI_ISL_622295\|2020 | .................... | .................... | ..................... | .................... | .................... | .................... |
| P | 4272 hCoV-19/Denmark/DCGC-6340/2020\|EPI_ISL_622296\|2020 | .................... | .................... | ..................... | .................... | .................... | .................... |
| P | 4273 hCoV-19/Denmark/DCGC-6346/2020\|EPI_ISL_622297\|2020 | .................... | .................... | ..................... | .................... | .................... | .................... |
| P | 4274 hCoV-19/Denmark/DCGC-6353/2020\|EPI_ISL_622298\|2020 | .................... | .................... | ..................... | .................... | .................... | .................... |
| P | 4275 hCoV-19/Denmark/DCGC-6364/2020\|EPI_ISL_622299\|2020 | .................... | .................... | ..................... | .................... | .................... | .................... |
| P | 4276 hCoV-19/Denmark/DCGC-6369/2020\|EPI_ISL_622300\|2020 | .................... | .................... | ..................... | .................... | .................... | .................... |
| P | 4277 hCoV-19/Denmark/DCGC-6382/2020\|EPI_ISL_622301\|2020 | .................... | .................... | ..................... | .................... | .................... | .................... |

| | | | | | | | | |
|---|---|---|---|---|---|---|---|---|
| P | 4278 | hCoV-19/Denmark/DCGC-6391/2020\|EPI_ISL_622302\|2020 | ···················· | ···················· | ····················· | ···················· | ···················· | ···················· |
| P | 4279 | hCoV-19/Denmark/DCGC-6347/2020\|EPI_ISL_622303\|2020 | ···················· | ···················· | ····················· | ···················· | ···················· | ···················· |
| P | 4280 | hCoV-19/Denmark/DCGC-6354/2020\|EPI_ISL_622304\|2020 | ···················· | ···················· | ····················· | ···················· | ···················· | ···················· |
| P | 4281 | hCoV-19/Denmark/DCGC-6402/2020\|EPI_ISL_622316\|2020 | ···················· | ···················· | ····················· | ···················· | ···················· | ···················· |
| P | 4282 | hCoV-19/Denmark/DCGC-6342/2020\|EPI_ISL_622317\|2020 | ···················· | ···················· | ····················· | ···················· | ···················· | ···················· |
| P | 4283 | hCoV-19/Denmark/DCGC-6372/2020\|EPI_ISL_622320\|2020 | ···················· | ···················· | ····················· | ···················· | ···················· | ···················· |
| P | 4284 | hCoV-19/Denmark/DCGC-6378/2020\|EPI_ISL_622321\|2020 | ···················· | ···················· | ····················· | ···················· | ···················· | ···················· |
| P | 4285 | hCoV-19/Denmark/DCGC-6385/2020\|EPI_ISL_622322\|2020 | ···················· | ···················· | ····················· | ···················· | ···················· | ···················· |
| P | 4286 | hCoV-19/Denmark/DCGC-6393/2020\|EPI_ISL_622323\|2020 | ···················· | ···················· | ····················· | ···················· | ···················· | ···················· |
| P | 4287 | hCoV-19/Denmark/DCGC-6398/2020\|EPI_ISL_622324\|2020 | ···················· | ·A·················· | ····················· | ···················· | ···················· | ···················· |
| P | 4288 | hCoV-19/Denmark/DCGC-6343/2020\|EPI_ISL_622325\|2020 | ···················· | ···················· | ····················· | ···················· | ···················· | ···················· |
| P | 4289 | hCoV-19/Denmark/DCGC-6350/2020\|EPI_ISL_622326\|2020 | ···················· | ···················· | ····················· | ···················· | ···················· | ···················· |
| P | 4290 | hCoV-19/Denmark/DCGC-6355/2020\|EPI_ISL_622327\|2020 | ···················· | ···················· | ····················· | ···················· | ···················· | ···················· |
| P | 4291 | hCoV-19/Denmark/DCGC-6360/2020\|EPI_ISL_622328\|2020 | ···················· | ···················· | ····················· | ···················· | ···················· | ···················· |
| P | 4292 | hCoV-19/Denmark/DCGC-6366/2020\|EPI_ISL_622329\|2020 | ···················· | ···················· | ····················· | ···················· | ···················· | ···················· |
| P | 4293 | hCoV-19/Denmark/DCGC-6373/2020\|EPI_ISL_622330\|2020 | ···················· | ···················· | ····················· | ···················· | ···················· | ···················· |
| P | 4294 | hCoV-19/Denmark/DCGC-6379/2020\|EPI_ISL_622331\|2020 | ···················· | ···················· | ····················· | ···················· | ···················· | ···················· |
| P | 4295 | hCoV-19/Denmark/DCGC-6387/2020\|EPI_ISL_622332\|2020 | ···················· | ···················· | ····················· | ···················· | ···················· | ···················· |
| P | 4296 | hCoV-19/Denmark/DCGC-6394/2020\|EPI_ISL_622333\|2020 | ···················· | ···················· | ····················· | ···················· | ···················· | ···················· |
| P | 4297 | hCoV-19/Denmark/DCGC-6344/2020\|EPI_ISL_622334\|2020 | ···················· | ···················· | ····················· | ···················· | ···················· | ···················· |
| P | 4298 | hCoV-19/Denmark/DCGC-6351/2020\|EPI_ISL_622335\|2020 | ···················· | ···················· | ····················· | ···················· | ···················· | ···················· |
| P | 4299 | hCoV-19/Denmark/DCGC-6361/2020\|EPI_ISL_622336\|2020 | ···················· | ···················· | ····················· | ···················· | ···················· | ···················· |
| P | 4300 | hCoV-19/Denmark/DCGC-6367/2020\|EPI_ISL_622337\|2020 | ···················· | ···················· | ····················· | ···················· | ···················· | ···················· |
| P | 4301 | hCoV-19/England/MILK-AB8D9D/2020\|EPI_ISL_623591\|20 | ···················· | ···················· | ····················· | ···················· | ···················· | ···················· |
| P | 4302 | hCoV-19/England/CAMC-AB14BD/2020\|EPI_ISL_623593\|20 | ···················· | ···················· | ····················· | ···················· | ···················· | ···················· |
| P | 4303 | hCoV-19/England/CAMC-AB2100/2020\|EPI_ISL_623595\|20 | ···················· | ···················· | ····················· | ···················· | ···················· | ···················· |
| P | 4304 | hCoV-19/England/MILK-AB7EF5/2020\|EPI_ISL_623596\|20 | ···················· | ···················· | ····················· | ···················· | ···················· | ···················· |
| P | 4305 | hCoV-19/England/MILK-AB85E0/2020\|EPI_ISL_623598\|20 | ···················· | ···················· | ····················· | ···················· | ···················· | ···················· |
| P | 4306 | hCoV-19/England/MILK-AB7C55/2020\|EPI_ISL_623599\|20 | ···················· | ···················· | ····················· | ···················· | ···················· | ···················· |
| P | 4307 | hCoV-19/England/MILK-AB7B3A/2020\|EPI_ISL_623631\|20 | ···················· | ···················· | ····················· | ···················· | ······T············· | ···················· |
| P | 4308 | hCoV-19/England/MILK-AB7EC8/2020\|EPI_ISL_623632\|20 | ···················· | ···················· | ····················· | ···················· | ···················· | ···················· |
| P | 4309 | hCoV-19/England/MILK-AB8E5E/2020\|EPI_ISL_623634\|20 | ···················· | ···················· | ····················· | ···················· | ···················· | ···················· |
| P | 4310 | hCoV-19/England/CAMC-AB1866/2020\|EPI_ISL_623647\|20 | ···················· | ···················· | ····················· | ···················· | ···················· | ···················· |
| P | 4311 | hCoV-19/England/MILK-AB8823/2020\|EPI_ISL_623648\|20 | ···················· | ···················· | ····················· | ···················· | ···················· | ···················· |
| P | 4312 | hCoV-19/England/MILK-AB87EA/2020\|EPI_ISL_623649\|20 | ···················· | ···················· | ····················· | ···················· | ···················· | ···················· |
| P | 4313 | hCoV-19/England/CAMC-AB1F16/2020\|EPI_ISL_623650\|20 | ···················· | ···················· | ····················· | ···················· | ···················· | ···················· |
| P | 4314 | hCoV-19/England/CAMC-AB1541/2020\|EPI_ISL_623651\|20 | ···················· | ···················· | ····················· | ···················· | ···················· | ···················· |
| P | 4315 | hCoV-19/England/CAMC-AB221C/2020\|EPI_ISL_623652\|20 | ···················· | ···················· | ····················· | ···················· | ···················· | ···················· |
| P | 4316 | hCoV-19/England/CAMC-AB15E7/2020\|EPI_ISL_623654\|20 | ···················· | ···················· | ····················· | ···················· | ···················· | ···················· |
| P | 4317 | hCoV-19/England/MILK-AB87AE/2020\|EPI_ISL_623656\|20 | ···················· | ···················· | ····················· | ···················· | ···················· | ···················· |
| P | 4318 | hCoV-19/England/CAMC-AB1DB2/2020\|EPI_ISL_623657\|20 | ···················· | ···················· | ····················· | ···················· | ···················· | ···················· |
| P | 4319 | hCoV-19/England/CAMC-AB18DF/2020\|EPI_ISL_623658\|20 | ···················· | ···················· | ····················· | ···················· | ···················· | ···················· |
| P | 4320 | hCoV-19/England/MILK-AB78CA/2020\|EPI_ISL_623659\|20 | ···················· | ···················· | ····················· | ···················· | ···················· | ···················· |
| P | 4321 | hCoV-19/England/MILK-AB861A/2020\|EPI_ISL_623660\|20 | ···················· | ···················· | ····················· | ···················· | ···················· | ···················· |
| P | 4322 | hCoV-19/England/CAMC-AB13A1/2020\|EPI_ISL_623662\|20 | ···················· | ···················· | ····················· | ···················· | ···················· | ···················· |
| P | 4323 | hCoV-19/England/CAMC-AB1954/2020\|EPI_ISL_623663\|20 | ···················· | ···················· | ····················· | ···················· | ···················· | ···················· |
| P | 4324 | hCoV-19/England/CAMC-AB1945/2020\|EPI_ISL_623665\|20 | ···················· | ···················· | ····················· | ···················· | ···················· | ···················· |
| P | 4325 | hCoV-19/England/CAMC-AB11B6/2020\|EPI_ISL_623666\|20 | ···················· | ···················· | ····················· | ···················· | ···················· | ···················· |
| P | 4326 | hCoV-19/England/CAMC-AB1D2B/2020\|EPI_ISL_623667\|20 | ···················· | ···················· | ····················· | ···················· | ···················· | ···················· |
| P | 4327 | hCoV-19/England/MILK-AB78D9/2020\|EPI_ISL_623668\|20 | ···················· | ···················· | ····················· | ···················· | ···················· | ···················· |
| P | 4328 | hCoV-19/England/ALDP-AA221F/2020\|EPI_ISL_624960\|20 | ···················· | ···················· | ····················· | ···················· | ···················· | ···················· |
| P | 4329 | hCoV-19/England/ALDP-AA0D2F/2020\|EPI_ISL_624961\|20 | ···················· | ···················· | ····················· | ···················· | ···················· | ···················· |
| P | 4330 | hCoV-19/England/ALDP-AA2CD5/2020\|EPI_ISL_624962\|20 | ···················· | ·A·················· | ····················· | ···················· | ···················· | ···················· |
| P | 4331 | hCoV-19/England/ALDP-AA3F71/2020\|EPI_ISL_624963\|20 | ···················· | ···················· | ····················· | ···················· | ···················· | ···················· |
| P | 4332 | hCoV-19/England/ALDP-AA0B61/2020\|EPI_ISL_624964\|20 | ···················· | ···················· | ····················· | ···················· | ···················· | ···················· |
| P | 4333 | hCoV-19/England/ALDP-AA5434/2020\|EPI_ISL_624965\|20 | ···················· | ···················· | ····················· | ···················· | ···················· | ···················· |
| P | 4334 | hCoV-19/England/ALDP-AA1122/2020\|EPI_ISL_624966\|20 | ···················· | ···················· | ····················· | ···················· | ···················· | ···················· |
| P | 4335 | hCoV-19/Wales/ALDP-AA3797/2020\|EPI_ISL_624978\|2020 | ···················· | ···················· | ····················· | ···················· | ···················· | ···················· |
| P | 4336 | hCoV-19/England/ALDP-AA0CE6/2020\|EPI_ISL_624980\|20 | ···················· | ···················· | ····················· | ···················· | ···················· | ···················· |
| P | 4337 | hCoV-19/England/ALDP-AA5759/2020\|EPI_ISL_624981\|20 | ···················· | ···················· | ····················· | ···················· | ···················· | ···················· |
| P | 4338 | hCoV-19/England/ALDP-AA36B8/2020\|EPI_ISL_624982\|20 | ···················· | ···················· | ····················· | ···················· | ···················· | ···················· |
| P | 4339 | hCoV-19/England/ALDP-AA33C0/2020\|EPI_ISL_624983\|20 | ···················· | ···················· | ····················· | ···················· | ···················· | ···················· |
| P | 4340 | hCoV-19/Wales/ALDP-AA38A3/2020\|EPI_ISL_624984\|2020 | ···················· | ···················· | ····················· | ···················· | ···················· | ···················· |
| P | 4341 | hCoV-19/England/ALDP-AA222E/2020\|EPI_ISL_624985\|20 | ···················· | ···················· | ····················· | ···················· | ···················· | ···················· |
| P | 4342 | hCoV-19/England/ALDP-AA16C9/2020\|EPI_ISL_624986\|20 | ···················· | ···················· | ····················· | ···················· | ···················· | ···················· |
| P | 4343 | hCoV-19/England/ALDP-AA0BF8/2020\|EPI_ISL_624988\|20 | ···················· | ···················· | ····················· | ···················· | ···················· | ···················· |
| P | 4344 | hCoV-19/England/ALDP-AA5601/2020\|EPI_ISL_624989\|20 | ···················· | ···················· | ····················· | ···················· | ···················· | ···················· |
| P | 4345 | hCoV-19/England/ALDP-AA085B/2020\|EPI_ISL_624990\|20 | ···················· | ···················· | ····················· | ···················· | ···················· | ···················· |
| P | 4346 | hCoV-19/England/ALDP-AA07D6/2020\|EPI_ISL_624991\|20 | ···················· | ···················· | ····················· | ···················· | ···················· | ···················· |
| P | 4347 | hCoV-19/England/ALDP-AA0C5F/2020\|EPI_ISL_624992\|20 | ···················· | ···················· | ····················· | ···················· | ···················· | ···················· |
| P | 4348 | hCoV-19/England/ALDP-AA0BBC/2020\|EPI_ISL_624994\|20 | ···················· | ···················· | ····················· | ···················· | ···················· | ···················· |
| P | 4349 | hCoV-19/England/ALDP-AA3EED/2020\|EPI_ISL_624996\|20 | ···················· | ···················· | ····················· | ···················· | ···················· | ···················· |
| P | 4350 | hCoV-19/England/ALDP-AA1E49/2020\|EPI_ISL_624997\|20 | ···················· | ···················· | ····················· | ···················· | ···················· | ···················· |
| P | 4351 | hCoV-19/England/ALDP-AA5777/2020\|EPI_ISL_624998\|20 | ···················· | ···················· | ····················· | ···················· | ···················· | ···················· |
| P | 4352 | hCoV-19/England/ALDP-AA1869/2020\|EPI_ISL_624999\|20 | ···················· | ···················· | ····················· | ···················· | ···················· | ···················· |
| P | 4353 | hCoV-19/Gibraltar/204462884/2020\|EPI_ISL_615051\|20 | ···················· | ···················· | ····················· | ···················· | ···················· | ···················· |
| P | 4354 | hCoV-19/Gibraltar/204462888/2020\|EPI_ISL_615053\|20 | ···················· | ···················· | ····················· | ···················· | ···················· | ···················· |
| P | 4355 | hCoV-19/Gibraltar/204462889/2020\|EPI_ISL_615054\|20 | ···················· | ···················· | ····················· | ···················· | ···················· | ···················· |
| P | 4356 | hCoV-19/Gibraltar/204462890/2020\|EPI_ISL_615055\|20 | ···················· | ···················· | ····················· | ···················· | ···················· | ···················· |
| P | 4357 | hCoV-19/Gibraltar/204462891/2020\|EPI_ISL_615056\|20 | ···················· | ···················· | ····················· | ···················· | ···················· | ···················· |

| | | | | | | | | |
|---|---|---|---|---|---|---|---|---|
| P | 4358 hCoV-19/Gibraltar/204462892/2020\|EPI_ISL_615057\|20 | ···················· | ···················· | ····················· | ···················· | ···················· | ···················· | |
| P | 4359 hCoV-19/Gibraltar/204462894/2020\|EPI_ISL_615058\|20 | ···················· | ···················· | ····················· | ···················· | ······T············· | ···················· | |
| P | 4360 hCoV-19/Gibraltar/204462895/2020\|EPI_ISL_615059\|20 | ···················· | ···················· | ····················· | ···················· | ···················· | ···················· | |
| P | 4361 hCoV-19/Gibraltar/204462897/2020\|EPI_ISL_615061\|20 | ···················· | ···················· | ····················· | ···················· | ···················· | ···················· | |
| P | 4362 hCoV-19/Gibraltar/204462898/2020\|EPI_ISL_615062\|20 | ···················· | ···················· | ····················· | ···················· | ···················· | ···················· | |
| P | 4363 hCoV-19/Sweden/20-21494/2020\|EPI_ISL_615076\|2020-0 | ···················· | ···················· | ····················· | ···················· | ···················· | ···················· | |
| P | 4364 hCoV-19/Sweden/20-21495/2020\|EPI_ISL_615077\|2020-0 | ···················· | ···················· | ····················· | ···················· | ···················· | ···················· | |
| P | 4365 hCoV-19/Sweden/20-21496/2020\|EPI_ISL_615078\|2020-0 | ···················· | ···················· | ····················· | ···················· | ···················· | ···················· | |
| P | 4366 hCoV-19/Sweden/20-22106/2020\|EPI_ISL_615079\|2020-0 | ···················· | ···················· | ····················· | ···················· | ···················· | ···················· | |
| P | 4367 hCoV-19/Sweden/20-22137/2020\|EPI_ISL_615080\|2020-0 | ···················· | ···················· | ····················· | ···················· | ···················· | ···················· | |
| P | 4368 hCoV-19/Sweden/20-22139/2020\|EPI_ISL_615081\|2020-0 | ···················· | ···················· | ····················· | ···················· | ···················· | ···················· | |
| P | 4369 hCoV-19/Sweden/20-22141/2020\|EPI_ISL_615082\|2020-0 | ···················· | ···················· | ····················· | ···················· | ···················· | ···················· | |
| P | 4370 hCoV-19/Sweden/20-22142/2020\|EPI_ISL_615083\|2020-0 | ···················· | ···················· | ····················· | ···················· | ···················· | ···················· | |
| P | 4371 hCoV-19/Sweden/20-22145/2020\|EPI_ISL_615084\|2020-0 | ···················· | ···················· | ····················· | ···················· | ···················· | ···················· | |
| P | 4372 hCoV-19/Sweden/20-22146/2020\|EPI_ISL_615085\|2020-0 | ···················· | ···················· | ····················· | ···················· | ···················· | ···················· | |
| P | 4373 hCoV-19/Sweden/20-22736/2020\|EPI_ISL_615086\|2020-0 | ···················· | ···················· | ····················· | ···················· | ···················· | ···················· | |
| P | 4374 hCoV-19/Sweden/20-51687/2020\|EPI_ISL_615087\|2020-0 | ···················· | ···················· | ····················· | ···················· | ···················· | ···················· | |
| P | 4375 hCoV-19/Sweden/20-51695/2020\|EPI_ISL_615088\|2020-0 | ···················· | ···················· | ····················· | ···················· | ···················· | ···················· | |
| P | 4376 hCoV-19/Sweden/20-51696/2020\|EPI_ISL_615089\|2020-0 | ···················· | ···················· | ····················· | ···················· | ···················· | ···················· | |
| P | 4377 hCoV-19/Sweden/20-51741/2020\|EPI_ISL_615090\|2020-0 | ···················· | ···················· | ····················· | ···················· | ···················· | ···················· | |
| P | 4378 hCoV-19/Sweden/20-51747/2020\|EPI_ISL_615091\|2020-0 | ···················· | ···················· | ··················T·· | ···················· | ···················· | ···················· | 1 |
| P | 4379 hCoV-19/Sweden/20-52111/2020\|EPI_ISL_615092\|2020-0 | ···················· | ···················· | ····················· | ···················· | ···················· | ···················· | |
| P | 4380 hCoV-19/Sweden/20-52112/2020\|EPI_ISL_615093\|2020-0 | ···················· | ···················· | ····················· | ···················· | ···················· | ···················· | |
| P | 4381 hCoV-19/Sweden/20-52114/2020\|EPI_ISL_615094\|2020-0 | ···················· | ···················· | ····················· | ···················· | ···················· | ···················· | |
| P | 4382 hCoV-19/Sweden/20-52116/2020\|EPI_ISL_615095\|2020-0 | ···················· | ···················· | ····················· | ···················· | ···················· | ···················· | |
| P | 4383 hCoV-19/Sweden/20-52118/2020\|EPI_ISL_615096\|2020-0 | ···················· | ···················· | ····················· | ···················· | ···················· | ···················· | |
| P | 4384 hCoV-19/Sweden/20-52694/2020\|EPI_ISL_615097\|2020-0 | ···················· | ···················· | ····················· | ···················· | ···················· | ···················· | |
| P | 4385 hCoV-19/Sweden/20-52699/2020\|EPI_ISL_615098\|2020-0 | ···················· | ···················· | ····················· | ···················· | ···················· | ···················· | |
| P | 4386 hCoV-19/Sweden/20-52700/2020\|EPI_ISL_615099\|2020-0 | ···················· | ···················· | ····················· | ···················· | ···················· | ···················· | |
| P | 4387 hCoV-19/Sweden/20-52702/2020\|EPI_ISL_615100\|2020-0 | ···················· | ···················· | ····················· | ···················· | ···················· | ···················· | |
| P | 4388 hCoV-19/Sweden/20-52697/2020\|EPI_ISL_615101\|2020-0 | ···················· | ···················· | ····················· | ···················· | ···················· | ···················· | |
| P | 4389 hCoV-19/Sweden/20-52698/2020\|EPI_ISL_615102\|2020-0 | ···················· | ···················· | ····················· | ···················· | ···················· | ···················· | |
| P | 4390 hCoV-19/Sweden/20-52817/2020\|EPI_ISL_615103\|2020-0 | ···················· | ···················· | ····················· | ···················· | ···················· | ···················· | |
| P | 4391 hCoV-19/Sweden/20-52882/2020\|EPI_ISL_615104\|2020-0 | ···················· | ···················· | ····················· | ···················· | ···················· | ···················· | |
| P | 4392 hCoV-19/Sweden/20-52883/2020\|EPI_ISL_615105\|2020-0 | ···················· | ···················· | ····················· | ···················· | ···················· | ···················· | |
| P | 4393 hCoV-19/Sweden/20-52885/2020\|EPI_ISL_615106\|2020-0 | ···················· | ···················· | ····················· | ···················· | ···················· | ···················· | |
| P | 4394 hCoV-19/Sweden/20-52886/2020\|EPI_ISL_615107\|2020-0 | ···················· | ···················· | ····················· | ···················· | ···················· | ···················· | |
| P | 4395 hCoV-19/Sweden/20-52779/2020\|EPI_ISL_615108\|2020-0 | ···················· | ·A·················· | ····················· | ···················· | ···················· | ···················· | |
| P | 4396 hCoV-19/Sweden/20-52748/2020\|EPI_ISL_615109\|2020-0 | ···················· | ···················· | ····················· | ···················· | ···················· | ···················· | |
| P | 4397 hCoV-19/Sweden/20-08964/2020\|EPI_ISL_615110\|2020-0 | ···················· | ·A·················· | ····················· | ···················· | ···················· | ··A················· | |
| P | 4398 hCoV-19/Sweden/20-52709/2020\|EPI_ISL_615111\|2020-0 | ···················· | ···················· | ····················· | ···················· | ···················· | ···················· | |
| P | 4399 hCoV-19/Sweden/20-52714/2020\|EPI_ISL_615112\|2020-0 | ···················· | ···················· | ····················· | ···················· | ···················· | ···················· | |
| P | 4400 hCoV-19/Sweden/20-52715/2020\|EPI_ISL_615113\|2020-0 | ···················· | ···················· | ····················· | ···················· | ···················· | ···················· | |
| P | 4401 hCoV-19/Sweden/20-52719/2020\|EPI_ISL_615114\|2020-0 | ···················· | ···················· | ····················· | ···················· | ···················· | ···················· | |
| P | 4402 hCoV-19/Sweden/20-52829/2020\|EPI_ISL_615115\|2020-0 | ···················· | ···················· | ····················· | ···················· | ···················· | ···················· | |
| P | 4403 hCoV-19/Sweden/20-52830/2020\|EPI_ISL_615116\|2020-0 | ···················· | ··G················· | ····················· | ···················· | ···················· | ···················· | |
| P | 4404 hCoV-19/Sweden/20-53000/2020\|EPI_ISL_615117\|2020-0 | ···················· | ···················· | ····················· | ···················· | ···················· | ···················· | |
| P | 4405 hCoV-19/Sweden/20-53001/2020\|EPI_ISL_615118\|2020-0 | ···················· | ···················· | ····················· | ···················· | ···················· | ···················· | |
| P | 4406 hCoV-19/Sweden/20-53003/2020\|EPI_ISL_615119\|2020-0 | ···················· | ···················· | ····················· | ···················· | ···················· | ···················· | |
| P | 4407 hCoV-19/Sweden/20-53007/2020\|EPI_ISL_615120\|2020-0 | ···················· | ···················· | ····················· | ···················· | ···················· | ···················· | |
| P | 4408 hCoV-19/Argentina/Heritas-HG023/2020\|EPI_ISL_61512 | ···················· | ···················· | ····················· | ··············A····· | ···················· | ···················· | |
| P | 4409 hCoV-19/Denmark/DCGC-6299/2020\|EPI_ISL_616380\|2020 | ···················· | ···················· | ····················· | ···················· | ···················· | ···················· | |
| P | 4410 hCoV-19/Denmark/DCGC-3613/2020\|EPI_ISL_616381\|2020 | ···················· | ···················· | ····················· | ···················· | ···················· | ···················· | |
| P | 4411 hCoV-19/Denmark/DCGC-3614/2020\|EPI_ISL_616382\|2020 | ···················· | ···················· | ····················· | ···················· | ···················· | ···················· | |
| P | 4412 hCoV-19/Denmark/DCGC-3615/2020\|EPI_ISL_616383\|2020 | ···················· | ···················· | ····················· | ···················· | ···················· | ···················· | |
| P | 4413 hCoV-19/Denmark/DCGC-3616/2020\|EPI_ISL_616384\|2020 | ···················· | ···················· | ····················· | ···················· | ···················· | ···················· | |
| P | 4414 hCoV-19/Denmark/DCGC-3617/2020\|EPI_ISL_616385\|2020 | ········T··········· | ···················· | ····················· | ···················· | ···················· | ···················· | |
| P | 4415 hCoV-19/Denmark/DCGC-3618/2020\|EPI_ISL_616386\|2020 | ···················· | ···················· | ····················· | ···················· | ···················· | ···················· | |
| P | 4416 hCoV-19/Denmark/DCGC-3619/2020\|EPI_ISL_616387\|2020 | ···················· | ···················· | ····················· | ···················· | ···················· | ···················· | |
| P | 4417 hCoV-19/Denmark/DCGC-3620/2020\|EPI_ISL_616388\|2020 | ···················· | ···················· | ····················· | ···················· | ···················· | ···················· | |
| P | 4418 hCoV-19/Denmark/DCGC-3621/2020\|EPI_ISL_616389\|2020 | ········T··········· | ···················· | ····················· | ···················· | ···················· | ···················· | |
| P | 4419 hCoV-19/Denmark/DCGC-3622/2020\|EPI_ISL_616390\|2020 | ···················· | ···················· | ····················· | ···················· | ···················· | ···················· | |
| P | 4420 hCoV-19/Denmark/DCGC-3623/2020\|EPI_ISL_616391\|2020 | ···················· | ···················· | ····················· | ···················· | ···················· | ···················· | |
| P | 4421 hCoV-19/Denmark/DCGC-3624/2020\|EPI_ISL_616392\|2020 | ···················· | ···················· | ····················· | ···················· | ···················· | ···················· | |
| P | 4422 hCoV-19/Denmark/DCGC-3625/2020\|EPI_ISL_616393\|2020 | ···················· | ···················· | ····················· | ···················· | ···················· | ···················· | |
| P | 4423 hCoV-19/Denmark/DCGC-3626/2020\|EPI_ISL_616394\|2020 | ···················· | ···················· | ····················· | ···················· | ···················· | ···················· | |
| P | 4424 hCoV-19/Denmark/DCGC-3627/2020\|EPI_ISL_616395\|2020 | ···················· | ···················· | ····················· | ···················· | ···················· | ···················· | |
| P | 4425 hCoV-19/Denmark/DCGC-3628/2020\|EPI_ISL_616396\|2020 | ···················· | ···················· | ····················· | ···················· | ···················· | ···················· | |
| P | 4426 hCoV-19/Denmark/DCGC-3629/2020\|EPI_ISL_616397\|2020 | ···················· | ···················· | ····················· | ···················· | ···················· | ···················· | |
| P | 4427 hCoV-19/Denmark/DCGC-6300/2020\|EPI_ISL_616398\|2020 | ···················· | ···················· | ····················· | ···················· | ···················· | ···················· | |
| P | 4428 hCoV-19/Denmark/DCGC-6301/2020\|EPI_ISL_616399\|2020 | ···················· | ···················· | ····················· | ···················· | ···················· | ···················· | |
| P | 4429 hCoV-19/Denmark/DCGC-2572/2020\|EPI_ISL_616430\|2020 | ···················· | ···················· | ····················· | ···················· | ···················· | ···················· | |
| P | 4430 hCoV-19/Denmark/DCGC-2699/2020\|EPI_ISL_616432\|2020 | ···················· | ···················· | ····················· | ···················· | ···················· | ···················· | |
| P | 4431 hCoV-19/Denmark/DCGC-2701/2020\|EPI_ISL_616434\|2020 | ···················· | ···················· | ····················· | ···················· | ···················· | ···················· | |
| P | 4432 hCoV-19/Denmark/DCGC-2702/2020\|EPI_ISL_616435\|2020 | ···················· | ···················· | ····················· | ···················· | ···················· | ···················· | |
| P | 4433 hCoV-19/Denmark/DCGC-2703/2020\|EPI_ISL_616436\|2020 | ···················· | ···················· | ····················· | ···················· | ···················· | ···················· | |
| P | 4434 hCoV-19/Denmark/DCGC-2704/2020\|EPI_ISL_616437\|2020 | ···················· | ·A·················· | ····················· | ···················· | ···················· | ··A················· | |
| P | 4435 hCoV-19/Denmark/DCGC-2705/2020\|EPI_ISL_616438\|2020 | ···················· | ···················· | ····················· | ···················· | ···················· | ···················· | |
| P | 4436 hCoV-19/Denmark/DCGC-2706/2020\|EPI_ISL_616439\|2020 | ···················· | ·A·················· | ····················· | ···················· | ···················· | ···················· | |
| P | 4437 hCoV-19/Denmark/DCGC-2707/2020\|EPI_ISL_616440\|2020 | ···················· | ···················· | ····················· | ···················· | ···················· | ···················· | |

| | | | | | | | | |
|---|---|---|---|---|---|---|---|---|
| P | 4438 | hCoV-19/Denmark/DCGC-2708/2020\|EPI_ISL_616441\|2020 | ···················· | ···················· | ····················· | ···················· | ···················· | ···················· |
| P | 4439 | hCoV-19/Denmark/DCGC-2710/2020\|EPI_ISL_616443\|2020 | ···················· | ···················· | ····················· | ···················· | ···················· | ···················· |
| P | 4440 | hCoV-19/Denmark/DCGC-2711/2020\|EPI_ISL_616444\|2020 | ···················· | ···················· | ····················· | ···················· | ···················· | ···················· |
| P | 4441 | hCoV-19/Denmark/DCGC-2712/2020\|EPI_ISL_616445\|2020 | ···················· | ···················· | ····················· | ···················· | ···················· | ···················· |
| P | 4442 | hCoV-19/Denmark/DCGC-2713/2020\|EPI_ISL_616446\|2020 | ···················· | ···················· | ····················· | ···················· | ···················· | ···················· |
| P | 4443 | hCoV-19/Denmark/DCGC-2714/2020\|EPI_ISL_616447\|2020 | ···················· | ···················· | ····················· | ···················· | ···················· | ···················· |
| P | 4444 | hCoV-19/Denmark/DCGC-2715/2020\|EPI_ISL_616448\|2020 | ···················· | ···················· | ····················· | ···················· | ···················· | ···················· |
| P | 4445 | hCoV-19/Denmark/DCGC-2716/2020\|EPI_ISL_616449\|2020 | ···················· | ···················· | ····················· | ···················· | ···················· | ···················· |
| P | 4446 | hCoV-19/Denmark/DCGC-2717/2020\|EPI_ISL_616450\|2020 | ···················· | ···················· | ····················· | ···················· | ···················· | ···················· |
| P | 4447 | hCoV-19/Denmark/DCGC-2815/2020\|EPI_ISL_616451\|2020 | ···················· | ···················· | ····················· | ···················· | ···················· | ···················· |
| P | 4448 | hCoV-19/Denmark/DCGC-2816/2020\|EPI_ISL_616452\|2020 | ···················· | ···················· | ····················· | ···················· | ···················· | ···················· |
| P | 4449 | hCoV-19/Denmark/DCGC-2817/2020\|EPI_ISL_616453\|2020 | ···················· | ···················· | ····················· | ···················· | ···················· | ···················· |
| P | 4450 | hCoV-19/Denmark/DCGC-2818/2020\|EPI_ISL_616454\|2020 | ···················· | ···················· | ····················· | ···················· | ···················· | ···················· |
| P | 4451 | hCoV-19/Denmark/DCGC-2819/2020\|EPI_ISL_616455\|2020 | ···················· | ···················· | ····················· | ···················· | ···················· | ···················· |
| P | 4452 | hCoV-19/Denmark/DCGC-2820/2020\|EPI_ISL_616456\|2020 | ···················· | ···················· | ····················· | ···················· | ···················· | ···················· |
| P | 4453 | hCoV-19/Denmark/DCGC-2821/2020\|EPI_ISL_616457\|2020 | ···················· | ···················· | ····················· | ···················· | ···················· | ···················· |
| P | 4454 | hCoV-19/Denmark/DCGC-2822/2020\|EPI_ISL_616458\|2020 | ···················· | ···················· | ····················· | ···················· | ···················· | ···················· |
| P | 4455 | hCoV-19/Denmark/DCGC-2489/2020\|EPI_ISL_617760\|2020 | ···················· | ···················· | ····················· | ···················· | ···················· | ···················· |
| P | 4456 | hCoV-19/Denmark/DCGC-2490/2020\|EPI_ISL_617761\|2020 | ···················· | ···················· | ····················· | ···················· | ···················· | ···················· |
| P | 4457 | hCoV-19/Denmark/DCGC-2491/2020\|EPI_ISL_617762\|2020 | ···················· | ···················· | ····················· | ···················· | ···················· | ···················· |
| P | 4458 | hCoV-19/Denmark/DCGC-2492/2020\|EPI_ISL_617763\|2020 | ···················· | ···················· | ····················· | ···················· | ···················· | ···················· |
| P | 4459 | hCoV-19/Denmark/DCGC-2493/2020\|EPI_ISL_617764\|2020 | ···················· | ···················· | ····················· | ···················· | ···················· | ···················· |
| P | 4460 | hCoV-19/Denmark/DCGC-2494/2020\|EPI_ISL_617765\|2020 | ···················· | ···················· | ····················· | ···················· | ···················· | ···················· |
| P | 4461 | hCoV-19/Denmark/DCGC-2495/2020\|EPI_ISL_617766\|2020 | ···················· | ···················· | ····················· | ···················· | ···················· | ···················· |
| P | 4462 | hCoV-19/Denmark/DCGC-2496/2020\|EPI_ISL_617767\|2020 | ···················· | ···················· | ····················· | ···················· | ···················· | ···················· |
| P | 4463 | hCoV-19/Denmark/DCGC-2497/2020\|EPI_ISL_617768\|2020 | ···················· | ···················· | ····················· | ···················· | ···················· | ···················· |
| P | 4464 | hCoV-19/Denmark/DCGC-2498/2020\|EPI_ISL_617769\|2020 | ···················· | ···················· | ····················· | ···················· | ···················· | ···················· |
| P | 4465 | hCoV-19/Denmark/DCGC-2499/2020\|EPI_ISL_617770\|2020 | ···················· | ···················· | ····················· | ···················· | ···················· | ···················· |
| P | 4466 | hCoV-19/Denmark/DCGC-2500/2020\|EPI_ISL_617771\|2020 | ···················· | ·A·················· | ····················· | ···················· | ···················· | ···················· |
| P | 4467 | hCoV-19/Denmark/DCGC-2501/2020\|EPI_ISL_617772\|2020 | ···················· | ·A·················· | ····················· | ···················· | ···················· | ···················· |
| P | 4468 | hCoV-19/Denmark/DCGC-2502/2020\|EPI_ISL_617773\|2020 | ···················· | ···················· | ····················· | ···················· | ···················· | ···················· |
| P | 4469 | hCoV-19/Denmark/DCGC-2503/2020\|EPI_ISL_617774\|2020 | ···················· | ···················· | ····················· | ···················· | ···················· | ···················· |
| P | 4470 | hCoV-19/Denmark/DCGC-2504/2020\|EPI_ISL_617775\|2020 | ···················· | ···················· | ····················· | ···················· | ···················· | ···················· |
| P | 4471 | hCoV-19/Denmark/DCGC-2505/2020\|EPI_ISL_617776\|2020 | ···················· | ···················· | ····················· | ···················· | ···················· | ···················· |
| P | 4472 | hCoV-19/Denmark/DCGC-2506/2020\|EPI_ISL_617777\|2020 | ···················· | ···················· | ····················· | ···················· | ···················· | ···················· |
| P | 4473 | hCoV-19/Denmark/DCGC-2507/2020\|EPI_ISL_617778\|2020 | ···················· | ···················· | ····················· | ···················· | ···················· | ···················· |
| P | 4474 | hCoV-19/Denmark/DCGC-2508/2020\|EPI_ISL_617779\|2020 | ···················· | ···················· | ····················· | ···················· | ···················· | ···················· |
| P | 4475 | hCoV-19/Denmark/DCGC-2509/2020\|EPI_ISL_617780\|2020 | ···················· | ···················· | ···········T········· | ···················· | ···················· | ···················· |
| P | 4476 | hCoV-19/Denmark/DCGC-2566/2020\|EPI_ISL_617781\|2020 | ···················· | ···················· | ····················· | ···················· | ···················· | ···················· |
| P | 4477 | hCoV-19/Denmark/DCGC-2510/2020\|EPI_ISL_617782\|2020 | ···················· | ···················· | ····················· | ···················· | ···················· | ···················· |
| P | 4478 | hCoV-19/Denmark/DCGC-2511/2020\|EPI_ISL_617783\|2020 | ···················· | ···················· | ····················· | ···················· | ···················· | ···················· |
| P | 4479 | hCoV-19/Denmark/DCGC-2512/2020\|EPI_ISL_617784\|2020 | ···················· | ···················· | ····················· | ···················· | ···················· | ···················· |
| P | 4480 | hCoV-19/Denmark/DCGC-2513/2020\|EPI_ISL_617785\|2020 | ···················· | ·A·················· | ····················· | ···················· | ···················· | ···················· |
| P | 4481 | hCoV-19/Denmark/DCGC-2514/2020\|EPI_ISL_617786\|2020 | ···················· | ···················· | ····················· | ···················· | ···················· | ···················· |
| P | 4482 | hCoV-19/Denmark/DCGC-2515/2020\|EPI_ISL_617787\|2020 | ···················· | ···················· | ····················· | ···················· | ···················· | ···················· |
| P | 4483 | hCoV-19/Denmark/DCGC-2516/2020\|EPI_ISL_617788\|2020 | ···················· | ···················· | ····················· | ···················· | ···················· | ···················· |
| P | 4484 | hCoV-19/Denmark/DCGC-2517/2020\|EPI_ISL_617789\|2020 | ···················· | ···················· | ····················· | ···················· | ···················· | ···················· |
| P | 4485 | hCoV-19/Denmark/DCGC-1371/2020\|EPI_ISL_622217\|2020 | ···················· | ···················· | ····················· | ···················· | ···················· | ···················· |
| P | 4486 | hCoV-19/Denmark/DCGC-1372/2020\|EPI_ISL_622218\|2020 | ···················· | ···················· | ····················· | ···················· | ···················· | ···················· |
| P | 4487 | hCoV-19/Denmark/DCGC-1373/2020\|EPI_ISL_622220\|2020 | ···················· | ···················· | ····················· | ···················· | ···················· | ···················· |
| P | 4488 | hCoV-19/Denmark/DCGC-1374/2020\|EPI_ISL_622221\|2020 | ···················· | ···················· | ····················· | ···················· | ···················· | ···················· |
| P | 4489 | hCoV-19/Denmark/DCGC-1375/2020\|EPI_ISL_622222\|2020 | ···················· | ···················· | ····················· | ···················· | ···················· | ···················· |
| P | 4490 | hCoV-19/Denmark/DCGC-1366/2020\|EPI_ISL_622223\|2020 | ···················· | ···················· | ····················· | ···················· | ···················· | ···················· |
| P | 4491 | hCoV-19/Denmark/DCGC-1367/2020\|EPI_ISL_622224\|2020 | ···················· | ···················· | ····················· | ···················· | ···················· | ···················· |
| P | 4492 | hCoV-19/Denmark/DCGC-1368/2020\|EPI_ISL_622225\|2020 | ···················· | ···················· | ····················· | ···················· | ···················· | ···················· |
| P | 4493 | hCoV-19/Denmark/DCGC-1369/2020\|EPI_ISL_622226\|2020 | ···················· | ···················· | ····················· | ···················· | ···················· | ···················· |
| P | 4494 | hCoV-19/Denmark/DCGC-1378/2020\|EPI_ISL_622229\|2020 | ···················· | ···················· | ····················· | ···················· | ···················· | ···················· |
| P | 4495 | hCoV-19/Denmark/DCGC-1380/2020\|EPI_ISL_622231\|2020 | ···················· | ···················· | ····················· | ···················· | ···················· | ···················· |
| P | 4496 | hCoV-19/Denmark/DCGC-1381/2020\|EPI_ISL_622232\|2020 | ···················· | ···················· | ····················· | ···················· | ···················· | ···················· |
| P | 4497 | hCoV-19/Denmark/DCGC-1382/2020\|EPI_ISL_622233\|2020 | ···················· | ···················· | ····················· | ···················· | ···················· | ···················· |
| P | 4498 | hCoV-19/Denmark/DCGC-1454/2020\|EPI_ISL_622234\|2020 | ···················· | ···················· | ····················· | ···················· | ···················· | ···················· |
| P | 4499 | hCoV-19/Denmark/DCGC-1455/2020\|EPI_ISL_622235\|2020 | ···················· | ···················· | ····················· | ···················· | ···················· | ···················· |
| P | 4500 | hCoV-19/Denmark/DCGC-1383/2020\|EPI_ISL_622236\|2020 | ···················· | ···················· | ····················· | ···················· | ···················· | ···················· |
| P | 4501 | hCoV-19/Denmark/DCGC-1384/2020\|EPI_ISL_622237\|2020 | ···················· | ···················· | ····················· | ···················· | ···················· | ···················· |
| P | 4502 | hCoV-19/Denmark/DCGC-1385/2020\|EPI_ISL_622238\|2020 | ···················· | ···················· | ····················· | ···················· | ···················· | ···················· |
| P | 4503 | hCoV-19/Denmark/DCGC-1744/2020\|EPI_ISL_622239\|2020 | ···················· | ···················· | ····················· | ···················· | ···················· | ···················· |
| P | 4504 | hCoV-19/Denmark/DCGC-1736/2020\|EPI_ISL_622240\|2020 | ···················· | ···················· | ····················· | ···················· | ···················· | ···················· |
| P | 4505 | hCoV-19/Denmark/DCGC-1737/2020\|EPI_ISL_622241\|2020 | ···················· | ···················· | ····················· | ···················· | ···················· | ···················· |
| P | 4506 | hCoV-19/Denmark/DCGC-1738/2020\|EPI_ISL_622242\|2020 | ···················· | ···················· | ····················· | ···················· | ···················· | ···················· |
| P | 4507 | hCoV-19/Denmark/DCGC-4574/2020\|EPI_ISL_622243\|2020 | ···················· | ···················· | ····················· | ···················· | ···················· | ···················· |
| P | 4508 | hCoV-19/Denmark/DCGC-4575/2020\|EPI_ISL_622244\|2020 | ···················· | ···················· | ····················· | ···················· | ···················· | ···················· |
| P | 4509 | hCoV-19/Denmark/DCGC-4576/2020\|EPI_ISL_622245\|2020 | ···················· | ···················· | ····················· | ···················· | ···················· | ···················· |
| P | 4510 | hCoV-19/Denmark/DCGC-4577/2020\|EPI_ISL_622246\|2020 | ···················· | ···················· | ····················· | ···················· | ···················· | ···················· |
| P | 4511 | hCoV-19/Denmark/DCGC-4578/2020\|EPI_ISL_622247\|2020 | ···················· | ···················· | ····················· | ···················· | ···················· | ···················· |
| P | 4512 | hCoV-19/Denmark/DCGC-4579/2020\|EPI_ISL_622248\|2020 | ···················· | ···················· | ····················· | ···················· | ···················· | ···················· |
| P | 4513 | hCoV-19/Denmark/DCGC-4580/2020\|EPI_ISL_622249\|2020 | ···················· | ···················· | ····················· | ···················· | ···················· | ···················· |
| P | 4514 | hCoV-19/Denmark/DCGC-4581/2020\|EPI_ISL_622250\|2020 | ···················· | ···················· | ····················· | ···················· | ···················· | ···················· |
| P | 4515 | hCoV-19/Denmark/DCGC-4582/2020\|EPI_ISL_622251\|2020 | ···················· | ···················· | ····················· | ···················· | ···················· | ···················· |
| P | 4516 | hCoV-19/Denmark/DCGC-4583/2020\|EPI_ISL_622252\|2020 | ···················· | ···················· | ····················· | ···················· | ···················· | ···················· |
| P | 4517 | hCoV-19/Denmark/DCGC-4584/2020\|EPI_ISL_622253\|2020 | ···················· | ···················· | ····················· | ···················· | ···················· | ···················· |

| | | | | | | | |
|---|---|---|---|---|---|---|---|
| P | 4518 hCoV-19/Denmark/DCGC-4585/2020\|EPI_ISL_622254\|2020 | ···················· | ···················· | ···················· | ···················· | ···················· | ···················· |
| P | 4519 hCoV-19/Denmark/DCGC-4586/2020\|EPI_ISL_622255\|2020 | ···················· | ···················· | ···················· | ···················· | ···················· | ···················· |
| P | 4520 hCoV-19/Denmark/DCGC-4588/2020\|EPI_ISL_622257\|2020 | ···················· | ···················· | ···················· | ···················· | ···················· | ···················· |
| P | 4521 hCoV-19/Denmark/DCGC-4589/2020\|EPI_ISL_622258\|2020 | ···················· | ···················· | ···················· | ···················· | ···················· | ···················· |
| P | 4522 hCoV-19/Denmark/DCGC-4590/2020\|EPI_ISL_622259\|2020 | ···················· | ···················· | ···················· | ···················· | ···················· | ···················· |
| P | 4523 hCoV-19/England/MILK-AB73C9/2020\|EPI_ISL_623500\|20 | ···················· | ···················· | ···················· | ···················· | ···················· | ···················· |
| P | 4524 hCoV-19/England/MILK-ABB5FC/2020\|EPI_ISL_623501\|20 | ···················· | ···················· | ···················· | ···················· | ···················· | ···················· |
| P | 4525 hCoV-19/England/MILK-AB8243/2020\|EPI_ISL_623502\|20 | ···················· | ···················· | ···················· | ···················· | ······T············· | ···············A···· |
| P | 4526 hCoV-19/England/MILK-AB7420/2020\|EPI_ISL_623503\|20 | ···················· | ···················· | ···················· | ···················· | ···················· | ···················· |
| P | 4527 hCoV-19/England/CAMC-AB211F/2020\|EPI_ISL_623548\|20 | ···················· | ···················· | ···················· | ···················· | ···················· | ···················· |
| P | 4528 hCoV-19/England/MILK-AB7CCE/2020\|EPI_ISL_623549\|20 | ···················· | ···················· | ···················· | ···················· | ···················· | ···················· |
| P | 4529 hCoV-19/England/CAMC-AB13B0/2020\|EPI_ISL_623550\|20 | ···················· | ···················· | ···················· | ···················· | ···················· | ···················· |
| P | 4530 hCoV-19/England/MILK-AB8744/2020\|EPI_ISL_623552\|20 | ···················· | ···················· | ···················· | ···················· | ····T··············· | ···················· |
| P | 4531 hCoV-19/England/MILK-AB8A96/2020\|EPI_ISL_623553\|20 | ···················· | ···················· | ···················· | ···················· | ···················· | ···················· |
| P | 4532 hCoV-19/England/CAMC-AB149F/2020\|EPI_ISL_623556\|20 | ···················· | ···················· | ···················· | ···················· | ···················· | ···················· |
| P | 4533 hCoV-19/England/CAMC-AB1909/2020\|EPI_ISL_623557\|20 | ···················· | ···················· | ···················· | ···················· | ···················· | ···················· |
| P | 4534 hCoV-19/England/CAMC-AB18A2/2020\|EPI_ISL_623559\|20 | ···················· | ···················· | ···················· | ···················· | ···················· | ···················· |
| P | 4535 hCoV-19/England/MILK-AB7B49/2020\|EPI_ISL_623561\|20 | ···················· | ···················· | ···················· | ···················· | ···················· | ···················· |
| P | 4536 hCoV-19/England/MILK-AB7578/2020\|EPI_ISL_623563\|20 | ···················· | ···················· | ···················· | ···················· | ····T··············· | ···················· |
| P | 4537 hCoV-19/England/MILK-AB7F3E/2020\|EPI_ISL_623564\|20 | ···················· | ···················· | ···················· | ···················· | ···················· | ···················· |
| P | 4538 hCoV-19/England/MILK-AB7BD0/2020\|EPI_ISL_623565\|20 | ···················· | ···················· | ···················· | ···················· | ···················· | ···················· |
| P | 4539 hCoV-19/England/MILK-AB7EE6/2020\|EPI_ISL_623566\|20 | ···················· | ···················· | ···················· | ···················· | ···················· | ···················· |
| P | 4540 hCoV-19/England/CAMC-AB19BE/2020\|EPI_ISL_623568\|20 | ···················· | ···················· | ···················· | ···················· | ···················· | ···················· |
| P | 4541 hCoV-19/England/CAMC-AB13FC/2020\|EPI_ISL_623569\|20 | ···················· | ···················· | ···················· | ···················· | ···················· | ···················· |
| P | 4542 hCoV-19/England/CAMC-AB1E73/2020\|EPI_ISL_623570\|20 | ···················· | ···················· | ···················· | ···················· | ···················· | ···················· |
| P | 4543 hCoV-19/England/CAMC-AB15BA/2020\|EPI_ISL_623572\|20 | ···················· | ···················· | ···················· | ···················· | ···················· | ···················· |
| P | 4544 hCoV-19/England/MILK-AB85FF/2020\|EPI_ISL_623573\|20 | ···················· | ···················· | ···················· | ···················· | ···················· | ···················· |
| P | 4545 hCoV-19/England/CAMC-AB1338/2020\|EPI_ISL_623574\|20 | ···················· | ···················· | ···················· | ···················· | ···················· | ···················· |
| P | 4546 hCoV-19/England/CAMC-AB1286/2020\|EPI_ISL_623575\|20 | ···················· | ···················· | ···················· | ···················· | ···················· | ···················· |
| P | 4547 hCoV-19/England/CAMC-AB13CF/2020\|EPI_ISL_623576\|20 | ···················· | ···················· | ···················· | ···················· | ···················· | ···················· |
| P | 4548 hCoV-19/England/MILK-AB9084/2020\|EPI_ISL_623577\|20 | ···················· | ···················· | ···················· | ···················· | ···················· | ···················· |
| P | 4549 hCoV-19/England/MILK-AB9057/2020\|EPI_ISL_623579\|20 | ···················· | ···················· | ···················· | ···················· | ···················· | ···················· |
| P | 4550 hCoV-19/England/MILK-AB7D34/2020\|EPI_ISL_623580\|20 | ···················· | ···················· | ···················· | ···················· | ···················· | ···················· |
| P | 4551 hCoV-19/England/MILK-AB7F10/2020\|EPI_ISL_623582\|20 | ···················· | ···················· | ···················· | ···················· | ···················· | ···················· |
| P | 4552 hCoV-19/England/MILK-AB7921/2020\|EPI_ISL_623583\|20 | ···················· | ···················· | ···················· | ···················· | ···················· | ···················· |
| P | 4553 hCoV-19/England/CAMC-AB2188/2020\|EPI_ISL_623584\|20 | ···················· | ···················· | ···················· | ···················· | ···················· | ···················· |
| P | 4554 hCoV-19/England/MILK-AB7F2F/2020\|EPI_ISL_623586\|20 | ···················· | ·A·················· | ···················· | ···················· | ···················· | ···A················ |
| P | 4555 hCoV-19/England/CAMC-AB1927/2020\|EPI_ISL_623587\|20 | ···················· | ···················· | ···················· | ···················· | ···················· | ···················· |
| P | 4556 hCoV-19/England/MILK-AB78F7/2020\|EPI_ISL_623588\|20 | ···················· | ···················· | ···················· | ···················· | ···················· | ···················· |
| P | 4557 hCoV-19/England/ALDP-A9474E/2020\|EPI_ISL_624800\|20 | ···················· | ···················· | ···················· | ···················· | ···················· | ···················· |
| P | 4558 hCoV-19/England/ALDP-A95640/2020\|EPI_ISL_624801\|20 | ···················· | ···················· | ···················· | ···················· | ···················· | ···················· |
| P | 4559 hCoV-19/England/ALDP-A95859/2020\|EPI_ISL_624802\|20 | ···················· | ···················· | ···················· | ···················· | ···················· | ···················· |
| P | 4560 hCoV-19/England/ALDP-A962A5/2020\|EPI_ISL_624803\|20 | ···················· | ···················· | ···················· | ···················· | ···················· | ···················· |
| P | 4561 hCoV-19/England/ALDP-A94A90/2020\|EPI_ISL_624804\|20 | ···················· | ···················· | ···················· | ···················· | ···················· | ···················· |
| P | 4562 hCoV-19/England/ALDP-A94623/2020\|EPI_ISL_624805\|20 | ···················· | ·A·················· | ···················· | ···················· | ···················· | ···A················ |
| P | 4563 hCoV-19/England/ALDP-A96515/2020\|EPI_ISL_624806\|20 | ···················· | ···················· | ···················· | ···················· | ···················· | ···················· |
| P | 4564 hCoV-19/England/ALDP-A9581D/2020\|EPI_ISL_624808\|20 | ···················· | ···················· | ···················· | ···················· | ···················· | ···················· |
| P | 4565 hCoV-19/England/ALDP-A95D4B/2020\|EPI_ISL_624809\|20 | ···················· | ···················· | ···················· | ···················· | ···················· | ···················· |
| P | 4566 hCoV-19/England/ALDP-A95FAF/2020\|EPI_ISL_624810\|20 | ···················· | ···················· | ···················· | ···················· | ···················· | ···················· |
| P | 4567 hCoV-19/England/ALDP-A96463/2020\|EPI_ISL_624811\|20 | ···················· | ···················· | ···················· | ···················· | ···················· | ···················· |
| P | 4568 hCoV-19/England/ALDP-A94447/2020\|EPI_ISL_624812\|20 | ···················· | ···················· | ···················· | ···················· | ···················· | ···················· |
| P | 4569 hCoV-19/England/ALDP-A95288/2020\|EPI_ISL_624813\|20 | ···················· | ···················· | ···················· | ···················· | ···················· | ···················· |
| P | 4570 hCoV-19/England/ALDP-A95A44/2020\|EPI_ISL_624814\|20 | ···················· | ···················· | ···················· | ···················· | ···················· | ···················· |
| P | 4571 hCoV-19/England/ALDP-A94104/2020\|EPI_ISL_624815\|20 | ···················· | ···················· | ···················· | ···················· | ···················· | ···················· |
| P | 4572 hCoV-19/England/ALDP-A96278/2020\|EPI_ISL_624816\|20 | ···················· | ···················· | ···················· | ···················· | ···················· | ···················· |
| P | 4573 hCoV-19/England/ALDP-A962F0/2020\|EPI_ISL_624817\|20 | ···················· | ···················· | ···················· | ···················· | ···················· | ···················· |
| P | 4574 hCoV-19/England/ALDP-A941F5/2020\|EPI_ISL_624819\|20 | ···················· | ···················· | ···················· | ···················· | ···················· | ···················· |
| P | 4575 hCoV-19/England/ALDP-A958A4/2020\|EPI_ISL_624820\|20 | ···················· | ···················· | ···················· | ···················· | ···················· | ···················· |
| P | 4576 hCoV-19/England/ALDP-A962C3/2020\|EPI_ISL_624821\|20 | ···················· | ·A·················· | ···················· | ···················· | ···················· | ···A················ |
| P | 4577 hCoV-19/England/ALDP-A9583B/2020\|EPI_ISL_624823\|20 | ···················· | ···················· | ···················· | ···················· | ···················· | ···················· |
| P | 4578 hCoV-19/England/ALDP-A953A3/2020\|EPI_ISL_624824\|20 | ···················· | ···················· | ···················· | ···················· | ···················· | ···················· |
| P | 4579 hCoV-19/England/ALDP-A95297/2020\|EPI_ISL_624825\|20 | ···················· | ···················· | ···················· | ···················· | ···················· | ···················· |
| P | 4580 hCoV-19/England/ALDP-A946C9/2020\|EPI_ISL_624826\|20 | ···················· | ···················· | ···················· | ···················· | ···················· | ···················· |
| P | 4581 hCoV-19/England/ALDP-A96409/2020\|EPI_ISL_624827\|20 | ···················· | ···················· | ···················· | ···················· | ···················· | ···················· |
| P | 4582 hCoV-19/England/ALDP-A95CD5/2020\|EPI_ISL_624828\|20 | ···················· | ···················· | ···················· | ···················· | ···················· | ···················· |
| P | 4583 hCoV-19/England/ALDP-A94B15/2020\|EPI_ISL_624829\|20 | ···················· | ···················· | ···················· | ···················· | ···················· | ···················· |
| P | 4584 hCoV-19/England/ALDP-A96005/2020\|EPI_ISL_624830\|20 | ···················· | ···················· | ···················· | ···················· | ···················· | ···················· |
| P | 4585 hCoV-19/England/ALDP-A9533A/2020\|EPI_ISL_624831\|20 | ···················· | ···················· | ···················· | ···················· | ···················· | ···················· |
| P | 4586 hCoV-19/England/ALDP-A9524C/2020\|EPI_ISL_624832\|20 | ···················· | ···················· | ···················· | ···················· | ···················· | ···················· |
| P | 4587 hCoV-19/England/ALDP-A9625A/2020\|EPI_ISL_624833\|20 | ···················· | ···················· | ···················· | ···················· | ···················· | ···················· |
| P | 4588 hCoV-19/England/ALDP-A95C4E/2020\|EPI_ISL_624834\|20 | ···················· | ···················· | ···················· | ···················· | ···················· | ···················· |
| P | 4589 hCoV-19/England/ALDP-A946E7/2020\|EPI_ISL_624847\|20 | ···················· | ···················· | ···················· | ···················· | ···················· | ···················· |
| P | 4590 hCoV-19/England/ALDP-A9523D/2020\|EPI_ISL_624879\|20 | ···················· | ···················· | ···················· | ···················· | ···················· | ···················· |
| P | 4591 hCoV-19/England/ALDP-A95F27/2020\|EPI_ISL_624880\|20 | ···················· | ·A·················· | ···················· | ···················· | ···················· | ···················· |
| P | 4592 hCoV-19/England/ALDP-A953D0/2020\|EPI_ISL_624881\|20 | ···················· | ···················· | ···················· | ···················· | ···················· | ···················· |
| P | 4593 hCoV-19/England/ALDP-A94B60/2020\|EPI_ISL_624882\|20 | ···················· | ···················· | ···················· | ···················· | ···················· | ···················· |
| P | 4594 hCoV-19/England/ALDP-A94C6D/2020\|EPI_ISL_624883\|20 | ···················· | ···················· | ···················· | ···················· | ···················· | ···················· |
| P | 4595 hCoV-19/England/ALDP-A95929/2020\|EPI_ISL_624884\|20 | ···················· | ···················· | ···················· | ···················· | ···················· | ···················· |
| P | 4596 hCoV-19/England/ALDP-A95E39/2020\|EPI_ISL_624886\|20 | ···················· | ···················· | ···················· | ···················· | ···················· | ···················· |
| P | 4597 hCoV-19/England/ALDP-A94A81/2020\|EPI_ISL_624887\|20 | ···················· | ···················· | ···················· | ···················· | ···················· | ···················· |

| | | | | | | | | |
|---|---|---|---|---|---|---|---|---|
| P | 4598 | hCoV-19/England/ALDP-A9417D/2020\|EPI_ISL_624888\|20 | .................... | .................... | .................... | .................... | .................... | .................... |
| P | 4599 | hCoV-19/England/ALDP-A9476C/2020\|EPI_ISL_624889\|20 | .................... | .................... | .................... | .................... | .................... | .................... |
| P | 4600 | hCoV-19/England/ALDP-A94571/2020\|EPI_ISL_624890\|20 | .................... | .................... | .................... | .................... | .................... | .................... |
| P | 4601 | hCoV-19/England/ALDP-A95FFA/2020\|EPI_ISL_624891\|20 | .................... | .................... | .................... | .................... | .................... | .................... |
| P | 4602 | hCoV-19/England/ALDP-A957B6/2020\|EPI_ISL_624892\|20 | .................... | .................... | .................... | .................... | .................... | .................... |
| P | 4603 | hCoV-19/England/ALDP-A95613/2020\|EPI_ISL_624893\|20 | .................... | .................... | .................... | .................... | .................... | .................... |
| P | 4604 | hCoV-19/England/ALDP-A95D69/2020\|EPI_ISL_624894\|20 | .................... | .................... | .................... | .................... | .................... | .................... |
| P | 4605 | hCoV-19/England/ALDP-A94650/2020\|EPI_ISL_624895\|20 | .................... | .................... | .................... | .................... | .................... | .................... |
| P | 4606 | hCoV-19/England/ALDP-A94C4F/2020\|EPI_ISL_624896\|20 | .................... | .................... | .................... | .................... | .................... | .................... |
| P | 4607 | hCoV-19/England/ALDP-A9582C/2020\|EPI_ISL_624898\|20 | .................... | .................... | .................... | .................... | .................... | .................... |
| P | 4608 | hCoV-19/England/ALDP-A94CD6/2020\|EPI_ISL_624899\|20 | .................... | .................... | .................... | .................... | .................... | .................... |
| P | 4609 | hCoV-19/Denmark/DCGC-945/2020\|EPI_ISL_614517\|2020- | .................... | .................... | .................... | .................... | .................... | .................... |
| P | 4610 | hCoV-19/Denmark/DCGC-950/2020\|EPI_ISL_614518\|2020- | .................... | .................... | .................... | .................... | .................... | .................... |
| P | 4611 | hCoV-19/Denmark/DCGC-921/2020\|EPI_ISL_614519\|2020- | .................... | .................... | .................... | .................... | .................... | .................... |
| P | 4612 | hCoV-19/Denmark/DCGC-923/2020\|EPI_ISL_614520\|2020- | .................... | .................... | .................... | .................... | .................... | .................... |
| P | 4613 | hCoV-19/Denmark/DCGC-926/2020\|EPI_ISL_614521\|2020- | .................... | .................... | .................... | .................... | .................... | .................... |
| P | 4614 | hCoV-19/Denmark/DCGC-1029/2020\|EPI_ISL_614522\|2020 | .................... | .................... | .................... | .................... | .................... | .................... |
| P | 4615 | hCoV-19/Denmark/DCGC-936/2020\|EPI_ISL_614523\|2020- | .................... | .................... | .................... | .................... | .................... | .................... |
| P | 4616 | hCoV-19/Denmark/DCGC-943/2020\|EPI_ISL_614524\|2020- | .................... | .................... | .................... | .................... | .................... | .................... |
| P | 4617 | hCoV-19/Denmark/DCGC-946/2020\|EPI_ISL_614525\|2020- | .................... | .................... | .................... | .................... | .................... | .................... |
| P | 4618 | hCoV-19/Denmark/DCGC-951/2020\|EPI_ISL_614526\|2020- | .................... | .................... | .................... | .................... | .................... | .................... |
| P | 4619 | hCoV-19/Denmark/DCGC-924/2020\|EPI_ISL_614527\|2020- | .................... | .................... | .................... | .................... | .................... | .................... |
| P | 4620 | hCoV-19/Denmark/DCGC-930/2020\|EPI_ISL_614528\|2020- | .................... | .................... | .................... | .................... | .................... | .................... |
| P | 4621 | hCoV-19/Denmark/DCGC-934/2020\|EPI_ISL_614529\|2020- | .................... | .................... | .................... | .................... | .................... | .................... |
| P | 4622 | hCoV-19/Denmark/DCGC-937/2020\|EPI_ISL_614530\|2020- | .................... | .................... | .................... | .................... | .................... | .................... |
| P | 4623 | hCoV-19/Denmark/DCGC-941/2020\|EPI_ISL_614531\|2020- | .................... | .................... | .................... | .................... | .................... | .................... |
| P | 4624 | hCoV-19/Denmark/DCGC-944/2020\|EPI_ISL_614532\|2020- | .................... | .................... | .................... | .................... | .................... | .................... |
| P | 4625 | hCoV-19/Denmark/DCGC-952/2020\|EPI_ISL_614533\|2020- | .................... | .................... | .................... | .................... | .................... | .................... |
| P | 4626 | hCoV-19/Denmark/DCGC-922/2020\|EPI_ISL_614534\|2020- | .................... | .................... | .................... | .................... | .................... | .................... |
| P | 4627 | hCoV-19/Denmark/DCGC-925/2020\|EPI_ISL_614535\|2020- | .................... | .................... | .................... | .................... | .................... | .................... |
| P | 4628 | hCoV-19/Denmark/DCGC-927/2020\|EPI_ISL_614536\|2020- | .................... | .................... | .................... | .................... | .................... | .................... |
| P | 4629 | hCoV-19/Denmark/DCGC-938/2020\|EPI_ISL_614537\|2020- | .................... | .................... | .................... | .................... | .................... | .................... |
| P | 4630 | hCoV-19/Denmark/DCGC-777/2020\|EPI_ISL_614538\|2020- | .................... | .................... | .................... | .................... | .................... | .................... |
| P | 4631 | hCoV-19/Denmark/DCGC-778/2020\|EPI_ISL_614539\|2020- | .................... | .................... | .................... | .................... | .................... | .................... |
| P | 4632 | hCoV-19/Denmark/DCGC-770/2020\|EPI_ISL_614540\|2020- | .................... | .................... | .................... | .................... | .................... | .................... |
| P | 4633 | hCoV-19/Denmark/DCGC-1030/2020\|EPI_ISL_614541\|2020 | .................... | .................... | .................... | .................... | .................... | .................... |
| P | 4634 | hCoV-19/Denmark/DCGC-948/2020\|EPI_ISL_614542\|2020- | .................... | .................... | .................... | .................... | .................... | .................... |
| P | 4635 | hCoV-19/Denmark/DCGC-757/2020\|EPI_ISL_614543\|2020- | .................... | .................... | .................... | .................... | .................... | .................... |
| P | 4636 | hCoV-19/Denmark/DCGC-765/2020\|EPI_ISL_614544\|2020- | .................... | .................... | .................... | .................... | .................... | .................... |
| P | 4637 | hCoV-19/Denmark/DCGC-1031/2020\|EPI_ISL_614545\|2020 | .................... | .................... | .................... | .................... | .................... | .................... |
| P | 4638 | hCoV-19/Denmark/DCGC-1032/2020\|EPI_ISL_614546\|2020 | .................... | .................... | .................... | .................... | .................... | .................... |
| P | 4639 | hCoV-19/Denmark/DCGC-953/2020\|EPI_ISL_614547\|2020- | .................... | .................... | .................... | .................... | .................... | .................... |
| P | 4640 | hCoV-19/Denmark/DCGC-956/2020\|EPI_ISL_614548\|2020- | .................... | .................... | .................... | .................... | .................... | .................... |
| P | 4641 | hCoV-19/Denmark/DCGC-963/2020\|EPI_ISL_614549\|2020- | .................... | .................... | .................... | .................... | .................... | .................... |
| P | 4642 | hCoV-19/Denmark/DCGC-966/2020\|EPI_ISL_614550\|2020- | .................... | .................... | .................... | .................... | .................... | .................... |
| P | 4643 | hCoV-19/Denmark/DCGC-969/2020\|EPI_ISL_614551\|2020- | .................... | .................... | .................... | .................... | .................... | .................... |
| P | 4644 | hCoV-19/Denmark/DCGC-976/2020\|EPI_ISL_614552\|2020- | .................... | .................... | .................... | .................... | .................... | .................... |
| P | 4645 | hCoV-19/Denmark/DCGC-980/2020\|EPI_ISL_614553\|2020- | .................... | .................... | .................... | .................... | .................... | .................... |
| P | 4646 | hCoV-19/Denmark/DCGC-983/2020\|EPI_ISL_614554\|2020- | .................... | .................... | .................... | .................... | .................... | .................... |
| P | 4647 | hCoV-19/Denmark/DCGC-988/2020\|EPI_ISL_614555\|2020- | .................... | .................... | .................... | .................... | .................... | .................... |
| P | 4648 | hCoV-19/Denmark/DCGC-957/2020\|EPI_ISL_614556\|2020- | .................... | .................... | .................... | .................... | .................... | .................... |
| P | 4649 | hCoV-19/Denmark/DCGC-961/2020\|EPI_ISL_614557\|2020- | .................... | .................... | .................... | .................... | .................... | .................... |
| P | 4650 | hCoV-19/Denmark/DCGC-964/2020\|EPI_ISL_614558\|2020- | .................... | .................... | .................... | .................... | .................... | .................... |
| P | 4651 | hCoV-19/Denmark/DCGC-967/2020\|EPI_ISL_614559\|2020- | .................... | .................... | .................... | .................... | .................... | .................... |
| P | 4652 | hCoV-19/Denmark/DCGC-973/2020\|EPI_ISL_614560\|2020- | .................... | .................... | .................... | .................... | .................... | .................... |
| P | 4653 | hCoV-19/Denmark/DCGC-977/2020\|EPI_ISL_614561\|2020- | .................... | .................... | .................... | .................... | .................... | .................... |
| P | 4654 | hCoV-19/Denmark/DCGC-981/2020\|EPI_ISL_614562\|2020- | .................... | .................... | .................... | .................... | .................... | .................... |
| P | 4655 | hCoV-19/Denmark/DCGC-989/2020\|EPI_ISL_614563\|2020- | .................... | .................... | .................... | .................... | .................... | .................... |
| P | 4656 | hCoV-19/Denmark/DCGC-954/2020\|EPI_ISL_614564\|2020- | .................... | .................... | .................... | .................... | .................... | .................... |
| P | 4657 | hCoV-19/Denmark/DCGC-958/2020\|EPI_ISL_614565\|2020- | .................... | .................... | .................... | .................... | .................... | .................... |
| P | 4658 | hCoV-19/Denmark/DCGC-1033/2020\|EPI_ISL_614566\|2020 | .................... | .................... | .................... | .................... | .................... | .................... |
| P | 4659 | hCoV-19/Denmark/DCGC-970/2020\|EPI_ISL_614567\|2020- | .................... | .................... | .................... | .................... | .................... | .................... |
| P | 4660 | hCoV-19/Denmark/DCGC-974/2020\|EPI_ISL_614568\|2020- | .................... | .................... | .................... | .................... | .................... | .................... |
| P | 4661 | hCoV-19/Denmark/DCGC-1035/2020\|EPI_ISL_614569\|2020 | .................... | .................... | .................... | .................... | .................... | .................... |
| P | 4662 | hCoV-19/Denmark/DCGC-985/2020\|EPI_ISL_614570\|2020- | .................... | .................... | .................... | .................... | .................... | .................... |
| P | 4663 | hCoV-19/Denmark/DCGC-990/2020\|EPI_ISL_614571\|2020- | .................... | .................... | .................... | .................... | .................... | .................... |
| P | 4664 | hCoV-19/Denmark/DCGC-994/2020\|EPI_ISL_614572\|2020- | .................... | .................... | .................... | .................... | .................... | .................... |
| P | 4665 | hCoV-19/Denmark/DCGC-959/2020\|EPI_ISL_614573\|2020- | .................... | .................... | .................... | .................... | .................... | .................... |
| P | 4666 | hCoV-19/Denmark/DCGC-962/2020\|EPI_ISL_614574\|2020- | .................... | .................... | .................... | .................... | .................... | .................... |
| P | 4667 | hCoV-19/Denmark/DCGC-971/2020\|EPI_ISL_614575\|2020- | .................... | .................... | .................... | .................... | .................... | .................... |
| P | 4668 | hCoV-19/Denmark/DCGC-975/2020\|EPI_ISL_614576\|2020- | .................... | .................... | .................... | .................... | .................... | .................... |
| P | 4669 | hCoV-19/Denmark/DCGC-979/2020\|EPI_ISL_614577\|2020- | .................... | .................... | .................... | .................... | .................... | .................... |
| P | 4670 | hCoV-19/Denmark/DCGC-982/2020\|EPI_ISL_614578\|2020- | .................... | .................... | .................... | .................... | .................... | .................... |
| P | 4671 | hCoV-19/Denmark/DCGC-995/2020\|EPI_ISL_614579\|2020- | .................... | .................... | .................... | .................... | .................... | .................... |
| P | 4672 | hCoV-19/Denmark/DCGC-986/2020\|EPI_ISL_614580\|2020- | .................... | .................... | .................... | .................... | .................... | .................... |
| P | 4673 | hCoV-19/Denmark/DCGC-991/2020\|EPI_ISL_614581\|2020- | .................... | .................... | .................... | .................... | .................... | .................... |
| P | 4674 | hCoV-19/Denmark/DCGC-996/2020\|EPI_ISL_614582\|2020- | .................... | .................... | .................... | .................... | .................... | .................... |
| P | 4675 | hCoV-19/Denmark/DCGC-955/2020\|EPI_ISL_614583\|2020- | .................... | .................... | .................... | .................... | .................... | .................... |
| P | 4676 | hCoV-19/Denmark/DCGC-960/2020\|EPI_ISL_614584\|2020- | .................... | .................... | .................... | .................... | .................... | .................... |
| P | 4677 | hCoV-19/Denmark/DCGC-1034/2020\|EPI_ISL_614585\|2020 | .................... | .................... | .................... | .................... | .................... | .................... |

| | | | | | | | |
|---|---|---|---|---|---|---|---|
| P | 4678 hCoV-19/Denmark/DCGC-987/2020\|EPI_ISL_614586\|2020- | ···················· | ···················· | ····················· | ···················· | ···················· | ···················· |
| P | 4679 hCoV-19/Denmark/DCGC-992/2020\|EPI_ISL_614587\|2020- | ···················· | ···················· | ····················· | ···················· | ···················· | ···················· |
| P | 4680 hCoV-19/Denmark/DCGC-1037/2020\|EPI_ISL_614588\|2020 | ···················· | ···················· | ····················· | ···················· | ···················· | ···················· |
| P | 4681 hCoV-19/Denmark/DCGC-1100/2020\|EPI_ISL_614589\|2020 | ···················· | ···················· | ····················· | ···················· | ···················· | ···················· |
| P | 4682 hCoV-19/Denmark/DCGC-1114/2020\|EPI_ISL_614590\|2020 | ···················· | ···················· | ····················· | ···················· | ···················· | ···················· |
| P | 4683 hCoV-19/Denmark/DCGC-1120/2020\|EPI_ISL_614591\|2020 | ···················· | ···················· | ····················· | ···················· | ···················· | ···················· |
| P | 4684 hCoV-19/Denmark/DCGC-1131/2020\|EPI_ISL_614592\|2020 | ···················· | ···················· | ····················· | ···················· | ···················· | ···················· |
| P | 4685 hCoV-19/Denmark/DCGC-1090/2020\|EPI_ISL_614593\|2020 | ···················· | ···················· | ····················· | ···················· | ···················· | ···················· |
| P | 4686 hCoV-19/Denmark/DCGC-1094/2020\|EPI_ISL_614594\|2020 | ···················· | ···················· | ····················· | ···················· | ···················· | ···················· |
| P | 4687 hCoV-19/Denmark/DCGC-1101/2020\|EPI_ISL_614595\|2020 | ···················· | ···················· | ····················· | ···················· | ···················· | ···················· |
| P | 4688 hCoV-19/Denmark/DCGC-1115/2020\|EPI_ISL_614596\|2020 | ···················· | ···················· | ····················· | ···················· | ···················· | ···················· |
| P | 4689 hCoV-19/Denmark/DCGC-1123/2020\|EPI_ISL_614597\|2020 | ···················· | ···················· | ····················· | ···················· | ···················· | ···················· |
| P | 4690 hCoV-19/Denmark/DCGC-1132/2020\|EPI_ISL_614598\|2020 | ···················· | ···················· | ····················· | ···················· | ···················· | ···················· |
| P | 4691 hCoV-19/Denmark/DCGC-1091/2020\|EPI_ISL_614599\|2020 | ···················· | ···················· | ····················· | ···················· | ···················· | ···················· |
| P | 4692 hCoV-19/Denmark/DCGC-3937/2020\|EPI_ISL_615848\|2020 | ···················· | ···················· | ····················· | ···················· | ···················· | ···················· |
| P | 4693 hCoV-19/Denmark/DCGC-3938/2020\|EPI_ISL_615849\|2020 | ···················· | ···················· | ····················· | ···················· | ···················· | ···················· |
| P | 4694 hCoV-19/Denmark/DCGC-3939/2020\|EPI_ISL_615850\|2020 | ···················· | ···················· | ····················· | ···················· | ···················· | ···················· |
| P | 4695 hCoV-19/Denmark/DCGC-3940/2020\|EPI_ISL_615851\|2020 | ···················· | ···················· | ····················· | ···················· | ···················· | ···················· |
| P | 4696 hCoV-19/Denmark/DCGC-3941/2020\|EPI_ISL_615852\|2020 | ···················· | ···················· | ····················· | ···················· | ···················· | ···················· |
| P | 4697 hCoV-19/Denmark/DCGC-3942/2020\|EPI_ISL_615853\|2020 | ···················· | ···················· | ····················· | ···················· | ···················· | ···················· |
| P | 4698 hCoV-19/Denmark/DCGC-3943/2020\|EPI_ISL_615854\|2020 | ···················· | ···················· | ····················· | ···················· | ···················· | ···················· |
| P | 4699 hCoV-19/Denmark/DCGC-3944/2020\|EPI_ISL_615855\|2020 | ···················· | ···················· | ····················· | ···················· | ···················· | ···················· |
| P | 4700 hCoV-19/Denmark/DCGC-3945/2020\|EPI_ISL_615856\|2020 | ···················· | ···················· | ····················· | ···················· | ···················· | ···················· |
| P | 4701 hCoV-19/Denmark/DCGC-3946/2020\|EPI_ISL_615857\|2020 | ···················· | ···················· | ····················· | ···················· | ···················· | ···················· |
| P | 4702 hCoV-19/Denmark/DCGC-3947/2020\|EPI_ISL_615858\|2020 | ···················· | ···················· | ····················· | ···················· | ···················· | ···················· |
| P | 4703 hCoV-19/Denmark/DCGC-3949/2020\|EPI_ISL_615860\|2020 | ···················· | ···················· | ····················· | ···················· | ···················· | ···················· |
| P | 4704 hCoV-19/Denmark/DCGC-3950/2020\|EPI_ISL_615861\|2020 | ···················· | ···················· | ····················· | ···················· | ···················· | ···················· |
| P | 4705 hCoV-19/Denmark/DCGC-3951/2020\|EPI_ISL_615862\|2020 | ···················· | ···················· | ····················· | ···················· | ···················· | ···················· |
| P | 4706 hCoV-19/Denmark/DCGC-3952/2020\|EPI_ISL_615863\|2020 | ···················· | ···················· | ····················· | ···················· | ···················· | ···················· |
| P | 4707 hCoV-19/Denmark/DCGC-3953/2020\|EPI_ISL_615864\|2020 | ···················· | ···················· | ····················· | ···················· | ···················· | ···················· |
| P | 4708 hCoV-19/Denmark/DCGC-3954/2020\|EPI_ISL_615865\|2020 | ···················· | ···················· | ····················· | ···················· | ···················· | ···················· |
| P | 4709 hCoV-19/Denmark/DCGC-3955/2020\|EPI_ISL_615866\|2020 | ···················· | ···················· | ····················· | ···················· | ···················· | ···················· |
| P | 4710 hCoV-19/Denmark/DCGC-3956/2020\|EPI_ISL_615867\|2020 | ···················· | ···················· | ····················· | ···················· | ···················· | ···················· |
| P | 4711 hCoV-19/Denmark/DCGC-3957/2020\|EPI_ISL_615868\|2020 | ···················· | ···················· | ····················· | ···················· | ···················· | ···················· |
| P | 4712 hCoV-19/Denmark/DCGC-3958/2020\|EPI_ISL_615869\|2020 | ···················· | ···················· | ····················· | ···················· | ···················· | ···················· |
| P | 4713 hCoV-19/Denmark/DCGC-3959/2020\|EPI_ISL_615870\|2020 | ···················· | ···················· | ····················· | ···················· | ···················· | ···················· |
| P | 4714 hCoV-19/Denmark/DCGC-3960/2020\|EPI_ISL_615871\|2020 | ···················· | ···················· | ····················· | ···················· | ···················· | ···················· |
| P | 4715 hCoV-19/Denmark/DCGC-3961/2020\|EPI_ISL_615872\|2020 | ···················· | ···················· | ····················· | ···················· | ···················· | ···················· |
| P | 4716 hCoV-19/Denmark/DCGC-3962/2020\|EPI_ISL_615873\|2020 | ···················· | ···················· | ····················· | ···················· | ···················· | ···················· |
| P | 4717 hCoV-19/Denmark/DCGC-3963/2020\|EPI_ISL_615875\|2020 | ···················· | ···················· | ····················· | ···················· | ···················· | ···················· |
| P | 4718 hCoV-19/Denmark/DCGC-3964/2020\|EPI_ISL_615876\|2020 | ···················· | ···················· | ····················· | ···················· | ···················· | ···················· |
| P | 4719 hCoV-19/Denmark/DCGC-3966/2020\|EPI_ISL_615879\|2020 | ···················· | ···················· | ····················· | ···················· | ···················· | ···················· |
| P | 4720 hCoV-19/Denmark/DCGC-3967/2020\|EPI_ISL_615880\|2020 | ···················· | ···················· | ····················· | ···················· | ···················· | ···················· |
| P | 4721 hCoV-19/Denmark/DCGC-3968/2020\|EPI_ISL_615881\|2020 | ···················· | ·A·················· | ····················· | ···················· | ···················· | ··A················· |
| P | 4722 hCoV-19/Denmark/DCGC-3969/2020\|EPI_ISL_615882\|2020 | ···················· | ·A·················· | ····················· | ···················· | ···················· | ··A················· |
| P | 4723 hCoV-19/Denmark/DCGC-3970/2020\|EPI_ISL_615883\|2020 | ···················· | ·A·················· | ····················· | ···················· | ···················· | ··A················· |
| P | 4724 hCoV-19/Denmark/DCGC-3971/2020\|EPI_ISL_615884\|2020 | ···················· | ···················· | ····················· | ···················· | ···················· | ···················· |
| P | 4725 hCoV-19/Denmark/DCGC-3972/2020\|EPI_ISL_615885\|2020 | ···················· | ···················· | ····················· | ···················· | ···················· | ···················· |
| P | 4726 hCoV-19/Denmark/DCGC-3973/2020\|EPI_ISL_615886\|2020 | ···················· | ···················· | ····················· | ···················· | ···················· | ···················· |
| P | 4727 hCoV-19/Denmark/DCGC-3974/2020\|EPI_ISL_615887\|2020 | ···················· | ···················· | ····················· | ···················· | ···················· | ···················· |
| P | 4728 hCoV-19/Denmark/DCGC-3975/2020\|EPI_ISL_615888\|2020 | ···················· | ···················· | ····················· | ···················· | ···················· | ···················· |
| P | 4729 hCoV-19/Denmark/DCGC-3976/2020\|EPI_ISL_615889\|2020 | ···················· | ···················· | ····················· | ···················· | ···················· | ···················· |
| P | 4730 hCoV-19/Denmark/DCGC-3977/2020\|EPI_ISL_615890\|2020 | ···················· | ···················· | ····················· | ···················· | ···················· | ···················· |
| P | 4731 hCoV-19/Denmark/DCGC-3978/2020\|EPI_ISL_615891\|2020 | ···················· | ···················· | ····················· | ···················· | ···················· | ···················· |
| P | 4732 hCoV-19/Denmark/DCGC-3979/2020\|EPI_ISL_615892\|2020 | ···················· | ···················· | ····················· | ···················· | ···················· | ···················· |
| P | 4733 hCoV-19/Denmark/DCGC-3980/2020\|EPI_ISL_615893\|2020 | ···················· | ···················· | ····················· | ···················· | ···················· | ···················· |
| P | 4734 hCoV-19/Denmark/DCGC-3981/2020\|EPI_ISL_615894\|2020 | ···················· | ···················· | ····················· | ···················· | ···················· | ···················· |
| P | 4735 hCoV-19/Denmark/DCGC-3982/2020\|EPI_ISL_615895\|2020 | ···················· | ···················· | ····················· | ···················· | ···················· | ···················· |
| P | 4736 hCoV-19/Denmark/DCGC-6328/2020\|EPI_ISL_615896\|2020 | ···················· | ···················· | ····················· | ···················· | ···················· | ···················· |
| P | 4737 hCoV-19/Denmark/DCGC-6329/2020\|EPI_ISL_615897\|2020 | ···················· | ···················· | ····················· | ···················· | ···················· | ···················· |
| P | 4738 hCoV-19/Denmark/DCGC-5227/2020\|EPI_ISL_615898\|2020 | ···················· | ···················· | ····················· | ···················· | ···················· | ···················· |
| P | 4739 hCoV-19/Denmark/DCGC-5228/2020\|EPI_ISL_615899\|2020 | ···················· | ···················· | ····················· | ···················· | ···················· | ···················· |
| P | 4740 hCoV-19/Denmark/DCGC-4761/2020\|EPI_ISL_622200\|2020 | ···················· | ···················· | ····················· | ···················· | ···················· | ···················· |
| P | 4741 hCoV-19/Denmark/DCGC-1733/2020\|EPI_ISL_622201\|2020 | ···················· | ···················· | ····················· | ···················· | ···················· | ···················· |
| P | 4742 hCoV-19/Denmark/DCGC-1735/2020\|EPI_ISL_622202\|2020 | ···················· | ···················· | ····················· | ···················· | ···················· | ···················· |
| P | 4743 hCoV-19/Denmark/DCGC-1723/2020\|EPI_ISL_622204\|2020 | ···················· | ···················· | ····················· | ···················· | ···················· | ···················· |
| P | 4744 hCoV-19/Denmark/DCGC-1355/2020\|EPI_ISL_622205\|2020 | ···················· | ···················· | ····················· | ···················· | ···················· | ···················· |
| P | 4745 hCoV-19/Denmark/DCGC-1356/2020\|EPI_ISL_622206\|2020 | ···················· | ···················· | ····················· | ···················· | ···················· | ···················· |
| P | 4746 hCoV-19/Denmark/DCGC-1357/2020\|EPI_ISL_622207\|2020 | ···················· | ···················· | ····················· | ···················· | ···················· | ···················· |
| P | 4747 hCoV-19/Denmark/DCGC-1358/2020\|EPI_ISL_622208\|2020 | ···················· | ···················· | ····················· | ···················· | ···················· | ···················· |
| P | 4748 hCoV-19/Denmark/DCGC-1359/2020\|EPI_ISL_622209\|2020 | ···················· | ···················· | ····················· | ···················· | ···················· | ···················· |
| P | 4749 hCoV-19/Denmark/DCGC-1360/2020\|EPI_ISL_622210\|2020 | ···················· | ···················· | ····················· | ···················· | ···················· | ···················· |
| P | 4750 hCoV-19/Denmark/DCGC-1361/2020\|EPI_ISL_622211\|2020 | ···················· | ···················· | ····················· | ···················· | ···················· | ···················· |
| P | 4751 hCoV-19/Denmark/DCGC-1362/2020\|EPI_ISL_622212\|2020 | ···················· | ···················· | ····················· | ···················· | ···················· | ···················· |
| P | 4752 hCoV-19/Denmark/DCGC-1363/2020\|EPI_ISL_622213\|2020 | ···················· | ···················· | ····················· | ···················· | ···················· | ···················· |
| P | 4753 hCoV-19/Denmark/DCGC-1364/2020\|EPI_ISL_622214\|2020 | ···················· | ···················· | ····················· | ···················· | ···················· | ···················· |
| P | 4754 hCoV-19/Denmark/DCGC-1365/2020\|EPI_ISL_622215\|2020 | ···················· | ···················· | ····················· | ···················· | ···················· | ···················· |
| P | 4755 hCoV-19/England/MILK-ABB5DE/2020\|EPI_ISL_623504\|20 | ···················· | ···················· | ····················· | ···················· | ···················· | ···················· |
| P | 4756 hCoV-19/England/MILK-ABB107/2020\|EPI_ISL_623505\|20 | ···················· | ···················· | ····················· | ···················· | ···················· | ···················· |
| P | 4757 hCoV-19/England/MILK-AB7D9E/2020\|EPI_ISL_623507\|20 | ···················· | ·A·················· | ····················· | ···················· | ···················· | ··A················· |

| | | | | | | | | |
|---|---|---|---|---|---|---|---|---|
| P | 4758 | hCoV-19/England/MILK-ABB644/2020\|EPI_ISL_623509\|20 | ···················· | ···················· | ···················· | ···················· | ···················· | ···················· |
| P | 4759 | hCoV-19/England/MILK-AB83E6/2020\|EPI_ISL_623510\|20 | ···················· | ···················· | ···················· | ···················· | ···················· | ···················· |
| P | 4760 | hCoV-19/England/MILK-AB7684/2020\|EPI_ISL_623511\|20 | ···················· | ···················· | ···················· | ···················· | ···················· | ···················· |
| P | 4761 | hCoV-19/England/MILK-ABB583/2020\|EPI_ISL_623512\|20 | ···················· | ···················· | ···················· | ···················· | ···················· | ···················· |
| P | 4762 | hCoV-19/England/MILK-AB86FC/2020\|EPI_ISL_623513\|20 | ···················· | ···················· | ···················· | ···················· | ···················· | ···················· |
| P | 4763 | hCoV-19/England/MILK-ABB3C5/2020\|EPI_ISL_623514\|20 | ···················· | ···················· | ···················· | ···················· | ···················· | ···················· |
| P | 4764 | hCoV-19/England/MILK-ABB073/2020\|EPI_ISL_623516\|20 | ···················· | ···················· | ···················· | ···················· | ·····T·············· | ···················· |
| P | 4765 | hCoV-19/England/MILK-ABB802/2020\|EPI_ISL_623517\|20 | ···················· | ···················· | ···················· | ···················· | ···················· | ···················· |
| P | 4766 | hCoV-19/England/MILK-ABB653/2020\|EPI_ISL_623519\|20 | ···················· | ···················· | ···················· | ···················· | ···················· | ···················· |
| P | 4767 | hCoV-19/England/CAMC-AAFDE4/2020\|EPI_ISL_623520\|20 | ···················· | ···················· | ···················· | ···················· | ···················· | ···················· |
| P | 4768 | hCoV-19/England/MILK-AB8708/2020\|EPI_ISL_623521\|20 | ···················· | ···················· | ···················· | ···················· | ·····T·············· | ···················· |
| P | 4769 | hCoV-19/England/MILK-AB8225/2020\|EPI_ISL_623523\|20 | ···················· | ···················· | ···················· | ···················· | ·····T·············· | ···················· |
| P | 4770 | hCoV-19/England/MILK-ABB18F/2020\|EPI_ISL_623524\|20 | ···················· | ·A·················· | ···················· | ···················· | ···················· | ··A················· |
| P | 4771 | hCoV-19/England/MILK-AB80FE/2020\|EPI_ISL_623525\|20 | ···················· | ···················· | ···················· | ···················· | ···················· | ···················· |
| P | 4772 | hCoV-19/England/CAMC-AAFE5A/2020\|EPI_ISL_623527\|20 | ···················· | ···················· | ···················· | ···················· | ···················· | ···················· |
| P | 4773 | hCoV-19/England/MILK-ABB6BD/2020\|EPI_ISL_623529\|20 | ···················· | ···················· | ···················· | ···················· | ···················· | ···················· |
| P | 4774 | hCoV-19/England/CAMC-AAFA83/2020\|EPI_ISL_623531\|20 | ···················· | ···················· | ···················· | ···················· | ···················· | ···················· |
| P | 4775 | hCoV-19/England/MILK-ABB6CC/2020\|EPI_ISL_623534\|20 | ···················· | ·A·················· | ···················· | ··············A····· | ···················· | ···················· |
| P | 4776 | hCoV-19/England/MILK-AB82BC/2020\|EPI_ISL_623535\|20 | ···················· | ···················· | ···················· | ···················· | ···················· | ···················· |
| P | 4777 | hCoV-19/England/MILK-ABB7BA/2020\|EPI_ISL_623536\|20 | ···················· | ···················· | ···················· | ···················· | ···················· | ···················· |
| P | 4778 | hCoV-19/England/CAMC-AAFE96/2020\|EPI_ISL_623537\|20 | ···················· | ···················· | ···················· | ···················· | ···················· | ···················· |
| P | 4779 | hCoV-19/England/CAMC-AAFED2/2020\|EPI_ISL_623539\|20 | ···················· | ···················· | ···················· | ···················· | ···················· | ···················· |
| P | 4780 | hCoV-19/England/MILK-AB83C8/2020\|EPI_ISL_623540\|20 | ···················· | ···················· | ···················· | ···················· | ···················· | ···················· |
| P | 4781 | hCoV-19/England/MILK-AB844D/2020\|EPI_ISL_623541\|20 | ···················· | ···················· | ···················· | ···················· | ···················· | ···················· |
| P | 4782 | hCoV-19/England/MILK-ABBA84/2020\|EPI_ISL_623542\|20 | ···················· | ···················· | ···················· | ···················· | ···················· | ···················· |
| P | 4783 | hCoV-19/England/CAMC-AAFDF3/2020\|EPI_ISL_623545\|20 | ···················· | ···················· | ···················· | ···················· | ·····T·············· | ···················· |
| P | 4784 | hCoV-19/England/CAMC-AB1E55/2020\|EPI_ISL_623547\|20 | ···················· | ···················· | ···················· | ···················· | ···················· | ···················· |
| P | 4785 | hCoV-19/England/ALDP-A94EA3/2020\|EPI_ISL_624836\|20 | ···················· | ···················· | ···················· | ···················· | ···················· | ···················· |
| P | 4786 | hCoV-19/England/ALDP-A963FD/2020\|EPI_ISL_624837\|20 | ···················· | ···················· | ···················· | ···················· | ···················· | ···················· |
| P | 4787 | hCoV-19/England/ALDP-A960D8/2020\|EPI_ISL_624838\|20 | ···················· | ···················· | ···················· | ···················· | ···················· | ···················· |
| P | 4788 | hCoV-19/England/ALDP-A964EB/2020\|EPI_ISL_624839\|20 | ···················· | ···················· | ···················· | ···················· | ···················· | ···················· |
| P | 4789 | hCoV-19/England/ALDP-A94C8B/2020\|EPI_ISL_624840\|20 | ···················· | ···················· | ···················· | ···················· | ···················· | ···················· |
| P | 4790 | hCoV-19/England/ALDP-A94614/2020\|EPI_ISL_624841\|20 | ···················· | ···················· | ···················· | ···················· | ···················· | ···················· |
| P | 4791 | hCoV-19/Wales/ALDP-A9572F/2020\|EPI_ISL_624848\|2020 | ···················· | ···················· | ···················· | ···················· | ···················· | ···················· |
| P | 4792 | hCoV-19/England/ALDP-A9469C/2020\|EPI_ISL_624849\|20 | ···················· | ···················· | ···················· | ···················· | ···················· | ···················· |
| P | 4793 | hCoV-19/England/ALDP-A95EFD/2020\|EPI_ISL_624851\|20 | ···················· | ···················· | ···················· | ···················· | ···················· | ···················· |
| P | 4794 | hCoV-19/England/ALDP-A95EDF/2020\|EPI_ISL_624852\|20 | ···················· | ···················· | ···················· | ···················· | ···················· | ···················· |
| P | 4795 | hCoV-19/England/ALDP-A955F8/2020\|EPI_ISL_624853\|20 | ···················· | ···················· | ···················· | ···················· | ···················· | ···················· |
| P | 4796 | hCoV-19/England/ALDP-A95D87/2020\|EPI_ISL_624854\|20 | ···················· | ···················· | ···················· | ···················· | ···················· | ···················· |
| P | 4797 | hCoV-19/England/ALDP-A9415F/2020\|EPI_ISL_624855\|20 | ···················· | ···················· | ···················· | ···················· | ···················· | ··A················· |
| P | 4798 | hCoV-19/England/ALDP-A94C30/2020\|EPI_ISL_624856\|20 | ···················· | ···················· | ···················· | ···················· | ···················· | ···················· |
| P | 4799 | hCoV-19/England/ALDP-A94E1C/2020\|EPI_ISL_624858\|20 | ···················· | ···················· | ···················· | ···················· | ···················· | ···················· |
| P | 4800 | hCoV-19/England/ALDP-A94D1F/2020\|EPI_ISL_624859\|20 | ···················· | ···················· | ···················· | ···················· | ···················· | ···················· |
| P | 4801 | hCoV-19/England/ALDP-A9631B/2020\|EPI_ISL_624860\|20 | ···················· | ···················· | ···················· | ···················· | ·····T·············· | ···················· |
| P | 4802 | hCoV-19/England/ALDP-A95570/2020\|EPI_ISL_624861\|20 | ···················· | ···················· | ···················· | ···················· | ···················· | ···················· |
| P | 4803 | hCoV-19/England/ALDP-A96384/2020\|EPI_ISL_624862\|20 | ···················· | ···················· | ···················· | ···················· | ···················· | ···················· |
| P | 4804 | hCoV-19/England/ALDP-A96287/2020\|EPI_ISL_624863\|20 | ···················· | ···················· | ···················· | ···················· | ···················· | ···················· |
| P | 4805 | hCoV-19/England/ALDP-A940DA/2020\|EPI_ISL_624865\|20 | ···················· | ···················· | ···················· | ···················· | ···················· | ··A················· |
| P | 4806 | hCoV-19/England/ALDP-A957E3/2020\|EPI_ISL_624866\|20 | ···················· | ···················· | ···················· | ···················· | ···················· | ···················· |
| P | 4807 | hCoV-19/England/ALDP-A9531C/2020\|EPI_ISL_624867\|20 | ···················· | ···················· | ···················· | ···················· | ···················· | ···················· |
| P | 4808 | hCoV-19/England/ALDP-A95C3F/2020\|EPI_ISL_624868\|20 | ···················· | ···················· | ···················· | ···················· | ···················· | ···················· |
| P | 4809 | hCoV-19/England/ALDP-A958D1/2020\|EPI_ISL_624869\|20 | ···················· | ···················· | ···················· | ···················· | ···················· | ···················· |
| P | 4810 | hCoV-19/England/ALDP-A96393/2020\|EPI_ISL_624870\|20 | ···················· | ···················· | ···················· | ···················· | ···················· | ···················· |
| P | 4811 | hCoV-19/England/ALDP-A960BA/2020\|EPI_ISL_624872\|20 | ···················· | ···················· | ···················· | ···················· | ···················· | ···················· |
| P | 4812 | hCoV-19/Wales/ALDP-A95631/2020\|EPI_ISL_624873\|2020 | ···················· | ···················· | ···················· | ···················· | ···················· | ···················· |
| P | 4813 | hCoV-19/England/ALDP-A95F72/2020\|EPI_ISL_624874\|20 | ···················· | ···················· | ···················· | ···················· | ···················· | ···················· |
| P | 4814 | hCoV-19/England/ALDP-A95DC3/2020\|EPI_ISL_624875\|20 | ···················· | ···················· | ···················· | ···················· | ···················· | ···················· |
| P | 4815 | hCoV-19/England/ALDP-A9608D/2020\|EPI_ISL_624876\|20 | ···················· | ···················· | ···················· | ···················· | ···················· | ···················· |
| P | 4816 | hCoV-19/England/ALDP-A952A6/2020\|EPI_ISL_624877\|20 | ···················· | ···················· | ···················· | ···················· | ···················· | ···················· |
| P | 4817 | hCoV-19/England/ALDP-A96445/2020\|EPI_ISL_624878\|20 | ···················· | ···················· | ···················· | ···················· | ···················· | ···················· |
| P | 4818 | hCoV-19/England/ALDP-AA214F/2020\|EPI_ISL_625200\|20 | ···················· | ···················· | ···················· | ···················· | ···················· | ···················· |
| P | 4819 | hCoV-19/England/ALDP-AA4AE8/2020\|EPI_ISL_625201\|20 | ···················· | ···················· | ···················· | ···················· | ···················· | ···················· |
| P | 4820 | hCoV-19/England/ALDP-AA0A64/2020\|EPI_ISL_625202\|20 | ···················· | ···················· | ···················· | ···················· | ···················· | ···················· |
| P | 4821 | hCoV-19/Wales/ALDP-AA3849/2020\|EPI_ISL_625203\|2020 | ···················· | ···················· | ···················· | ···················· | ···················· | ···················· |
| P | 4822 | hCoV-19/England/ALDP-AA323C/2020\|EPI_ISL_625204\|20 | ···················· | ···················· | ···················· | ···················· | ···················· | ···················· |
| P | 4823 | hCoV-19/England/ALDP-AA166F/2020\|EPI_ISL_625205\|20 | ···················· | ···················· | ···················· | ···················· | ···················· | ···················· |
| P | 4824 | hCoV-19/England/ALDP-AA22A6/2020\|EPI_ISL_625206\|20 | ···················· | ···················· | ···················· | ···················· | ···················· | ···················· |
| P | 4825 | hCoV-19/England/ALDP-AA3F35/2020\|EPI_ISL_625207\|20 | ···················· | ···················· | ···················· | ···················· | ···················· | ···················· |
| P | 4826 | hCoV-19/England/ALDP-AA2C5D/2020\|EPI_ISL_625208\|20 | ···················· | ·A·················· | ···················· | ···················· | ···················· | ···················· |
| P | 4827 | hCoV-19/England/ALDP-AA2CF3/2020\|EPI_ISL_625209\|20 | ···················· | ···················· | ···················· | ···················· | ···················· | ···················· |
| P | 4828 | hCoV-19/England/ALDP-AA4990/2020\|EPI_ISL_625210\|20 | ···················· | ···················· | ···················· | ···················· | ···················· | ···················· |
| P | 4829 | hCoV-19/England/ALDP-AA17F3/2020\|EPI_ISL_625211\|20 | ···················· | ···················· | ···················· | ···················· | ···················· | ···················· |
| P | 4830 | hCoV-19/Wales/ALDP-AA5452/2020\|EPI_ISL_625212\|2020 | ···················· | ···················· | ···················· | ···················· | ···················· | ···················· |
| P | 4831 | hCoV-19/England/ALDP-AA1799/2020\|EPI_ISL_625214\|20 | ···················· | ···················· | ···················· | ···················· | ···················· | ···················· |
| P | 4832 | hCoV-19/England/ALDP-AA52A3/2020\|EPI_ISL_625215\|20 | ···················· | ···················· | ···················· | ···················· | ···················· | ···················· |
| P | 4833 | hCoV-19/England/ALDP-AA0CF5/2020\|EPI_ISL_625216\|20 | ···················· | ···················· | ···················· | ···················· | ···················· | ···················· |
| P | 4834 | hCoV-19/England/ALDP-AA54BC/2020\|EPI_ISL_625217\|20 | ···················· | ···················· | ···················· | ···················· | ···················· | ···················· |
| P | 4835 | hCoV-19/England/ALDP-AA2C6C/2020\|EPI_ISL_625218\|20 | ···················· | ···················· | ···················· | ···················· | ···················· | ···················· |
| P | 4836 | hCoV-19/England/ALDP-AA216D/2020\|EPI_ISL_625219\|20 | ···················· | ···················· | ···················· | ···················· | ···················· | ···················· |
| P | 4837 | hCoV-19/England/ALDP-AA2121/2020\|EPI_ISL_625220\|20 | ···················· | ·········A·········· | ···················· | ···················· | ···················· | ···················· |

| | | | | | | | | |
|---|---|---|---|---|---|---|---|---|
| P | 4838 | hCoV-19/England/ALDP-AA44DB/2020\|EPI_ISL_625221\|20 | ···················· | ···················· | ····················· | ···················· | ···················· | ···················· |
| P | 4839 | hCoV-19/England/ALDP-AA0AEC/2020\|EPI_ISL_625223\|20 | ···················· | ···················· | ····················· | ···················· | ···················· | ···················· |
| P | 4840 | hCoV-19/England/ALDP-AA1DE2/2020\|EPI_ISL_625224\|20 | ···················· | ···················· | ····················· | ···················· | ···················· | ···················· |
| P | 4841 | hCoV-19/England/ALDP-AA32C3/2020\|EPI_ISL_625226\|20 | ···················· | ···················· | ····················· | ···················· | ···················· | ···················· |
| P | 4842 | hCoV-19/England/ALDP-AA4550/2020\|EPI_ISL_625227\|20 | ···················· | ···················· | ···········T········· | ···················· | ···················· | ···················· |
| P | 4843 | hCoV-19/England/ALDP-AA3409/2020\|EPI_ISL_625228\|20 | ············T······· | ···················· | ····················· | ···················· | ···················· | ···················· |
| P | 4844 | hCoV-19/England/ALDP-AA177B/2020\|EPI_ISL_625229\|20 | ···················· | ···················· | ····················· | ···················· | ···················· | ···················· |
| P | 4845 | hCoV-19/England/ALDP-AA5364/2020\|EPI_ISL_625230\|20 | ···················· | ···················· | ····················· | ···················· | ···················· | ···················· |
| P | 4846 | hCoV-19/England/ALDP-AA180F/2020\|EPI_ISL_625231\|20 | ···················· | ···················· | ····················· | ···················· | ···················· | ···················· |
| P | 4847 | hCoV-19/England/ALDP-AA366D/2020\|EPI_ISL_625232\|20 | ···················· | ···················· | ····················· | ···················· | ···················· | ···················· |
| P | 4848 | hCoV-19/England/ALDP-AA53A0/2020\|EPI_ISL_625233\|20 | ···················· | ···················· | ····················· | ···················· | ···················· | ···················· |
| P | 4849 | hCoV-19/England/ALDP-AA53FB/2020\|EPI_ISL_625234\|20 | ···················· | ·A·················· | ····················· | ···················· | ···················· | ···················· |
| P | 4850 | hCoV-19/England/ALDP-AA2C3F/2020\|EPI_ISL_625236\|20 | ···················· | ·A·················· | ····················· | ···················· | ···················· | ···················· |
| P | 4851 | hCoV-19/England/ALDP-AA563E/2020\|EPI_ISL_625237\|20 | ···················· | ···················· | ····················· | ···················· | ···················· | ···················· |
| P | 4852 | hCoV-19/England/ALDP-AA084C/2020\|EPI_ISL_625238\|20 | ···················· | ·A·················· | ····················· | ···················· | ···················· | ···················· |
| P | 4853 | hCoV-19/England/ALDP-AA21A9/2020\|EPI_ISL_625239\|20 | ···················· | ···················· | ····················· | ···················· | ···················· | ···················· |
| P | 4854 | hCoV-19/England/ALDP-AA22C4/2020\|EPI_ISL_625241\|20 | ···················· | ···················· | ····················· | ···················· | ···················· | ···················· |
| P | 4855 | hCoV-19/England/ALDP-AA3885/2020\|EPI_ISL_625242\|20 | ···················· | ···················· | ····················· | ···················· | ···················· | ···················· |
| P | 4856 | hCoV-19/England/ALDP-AA5768/2020\|EPI_ISL_625243\|20 | ···················· | ···················· | ····················· | ···················· | ···················· | ···················· |
| P | 4857 | hCoV-19/England/ALDP-AA3F62/2020\|EPI_ISL_625244\|20 | ···················· | ···················· | ····················· | ···················· | ···················· | ···················· |
| P | 4858 | hCoV-19/England/ALDP-AA1113/2020\|EPI_ISL_625245\|20 | ···················· | ···················· | ····················· | ···················· | ···················· | ···················· |
| P | 4859 | hCoV-19/England/ALDP-AA0C6E/2020\|EPI_ISL_625246\|20 | ···················· | ···················· | ····················· | ···················· | ···················· | ···················· |
| P | 4860 | hCoV-19/England/ALDP-AA38B2/2020\|EPI_ISL_625248\|20 | ···················· | ···················· | ····················· | ···················· | ···················· | ···················· |
| P | 4861 | hCoV-19/England/ALDP-AA54CB/2020\|EPI_ISL_625249\|20 | ···················· | ···················· | ····················· | ···················· | ···················· | ···················· |
| P | 4862 | hCoV-19/Scotland/CAMC-A82057/2020\|EPI_ISL_625290\|2 | ···················· | ···················· | ····················· | ···················· | ···················· | ···················· |
| P | 4863 | hCoV-19/Wales/CAMC-A80E13/2020\|EPI_ISL_625291\|2020 | ···················· | ···················· | ····················· | ···················· | ···················· | ···················· |
| P | 4864 | hCoV-19/Scotland/CAMC-A81F88/2020\|EPI_ISL_625293\|2 | ···················· | ···················· | ····················· | ···················· | ···················· | ···················· |
| P | 4865 | hCoV-19/Denmark/DCGC-1327/2020\|EPI_ISL_618000\|2020 | ···················· | ···················· | ····················· | ···················· | ···················· | ···················· |
| P | 4866 | hCoV-19/Denmark/DCGC-1328/2020\|EPI_ISL_618001\|2020 | ···················· | ···················· | ····················· | ···················· | ···················· | ···················· |
| P | 4867 | hCoV-19/Denmark/DCGC-1329/2020\|EPI_ISL_618002\|2020 | ···················· | ···················· | ····················· | ···················· | ···················· | ···················· |
| P | 4868 | hCoV-19/Denmark/DCGC-1330/2020\|EPI_ISL_618003\|2020 | ···················· | ···················· | ····················· | ···················· | ···················· | ···················· |
| P | 4869 | hCoV-19/Denmark/DCGC-1331/2020\|EPI_ISL_618004\|2020 | ···················· | ···················· | ····················· | ···················· | ···················· | ···················· |
| P | 4870 | hCoV-19/Denmark/DCGC-1332/2020\|EPI_ISL_618005\|2020 | ···················· | ···················· | ····················· | ···················· | ···················· | ···················· |
| P | 4871 | hCoV-19/Denmark/DCGC-1453/2020\|EPI_ISL_618006\|2020 | ···················· | ···················· | ····················· | ···················· | ···················· | ···················· |
| P | 4872 | hCoV-19/Denmark/DCGC-1334/2020\|EPI_ISL_618008\|2020 | ···················· | ···················· | ····················· | ···················· | ···················· | ···················· |
| P | 4873 | hCoV-19/Denmark/DCGC-1335/2020\|EPI_ISL_618009\|2020 | ···················· | ···················· | ····················· | ···················· | ···················· | ···················· |
| P | 4874 | hCoV-19/Denmark/DCGC-1336/2020\|EPI_ISL_618010\|2020 | ···················· | ···················· | ····················· | ···················· | ···················· | ···················· |
| P | 4875 | hCoV-19/Denmark/DCGC-1337/2020\|EPI_ISL_618011\|2020 | ···················· | ···················· | ····················· | ···················· | ···················· | ···················· |
| P | 4876 | hCoV-19/Denmark/DCGC-1338/2020\|EPI_ISL_618012\|2020 | ···················· | ···················· | ····················· | ···················· | ···················· | ···················· |
| P | 4877 | hCoV-19/Denmark/DCGC-1259/2020\|EPI_ISL_618014\|2020 | ···················· | ···················· | ····················· | ···················· | ···················· | ···················· |
| P | 4878 | hCoV-19/Denmark/DCGC-1258/2020\|EPI_ISL_618015\|2020 | ···················· | ···················· | ····················· | ···················· | ···················· | ···················· |
| P | 4879 | hCoV-19/Denmark/DCGC-1257/2020\|EPI_ISL_618016\|2020 | ···················· | ···················· | ····················· | ···················· | ···················· | ···················· |
| P | 4880 | hCoV-19/Denmark/DCGC-1256/2020\|EPI_ISL_618017\|2020 | ···················· | ···················· | ····················· | ···················· | ···················· | ···················· |
| P | 4881 | hCoV-19/Denmark/DCGC-1254/2020\|EPI_ISL_618019\|2020 | ···················· | ···················· | ····················· | ···················· | ···················· | ···················· |
| P | 4882 | hCoV-19/Denmark/DCGC-1253/2020\|EPI_ISL_618020\|2020 | ···················· | ···················· | ····················· | ···················· | ···················· | ···················· |
| P | 4883 | hCoV-19/Denmark/DCGC-1252/2020\|EPI_ISL_618021\|2020 | ···················· | ···················· | ····················· | ···················· | ···················· | ···················· |
| P | 4884 | hCoV-19/Denmark/DCGC-1251/2020\|EPI_ISL_618022\|2020 | ···················· | ···················· | ····················· | ···················· | ···················· | ···················· |
| P | 4885 | hCoV-19/Denmark/DCGC-1261/2020\|EPI_ISL_618023\|2020 | ···················· | ···················· | ····················· | ···················· | ···················· | ···················· |
| P | 4886 | hCoV-19/Denmark/DCGC-1819/2020\|EPI_ISL_618024\|2020 | ···················· | ···················· | ····················· | ···················· | ···················· | ···················· |
| P | 4887 | hCoV-19/Denmark/DCGC-1350/2020\|EPI_ISL_618025\|2020 | ···················· | ···················· | ····················· | ···················· | ···················· | ···················· |
| P | 4888 | hCoV-19/Denmark/DCGC-1260/2020\|EPI_ISL_618026\|2020 | ···················· | ···················· | ····················· | ···················· | ···················· | ···················· |
| P | 4889 | hCoV-19/Denmark/DCGC-2070/2020\|EPI_ISL_618027\|2020 | ···················· | ···················· | ····················· | ···················· | ···················· | ···················· |
| P | 4890 | hCoV-19/Denmark/DCGC-2071/2020\|EPI_ISL_618028\|2020 | ···················· | ···················· | ····················· | ···················· | ···················· | ···················· |
| P | 4891 | hCoV-19/Denmark/DCGC-2072/2020\|EPI_ISL_618029\|2020 | ···················· | ···················· | ····················· | ···················· | ···················· | ···················· |
| P | 4892 | hCoV-19/Denmark/DCGC-2073/2020\|EPI_ISL_618030\|2020 | ···················· | ···················· | ····················· | ···················· | ···················· | ···················· |
| P | 4893 | hCoV-19/Denmark/DCGC-2074/2020\|EPI_ISL_618031\|2020 | ···················· | ···················· | ····················· | ···················· | ···················· | ···················· |
| P | 4894 | hCoV-19/Denmark/DCGC-1822/2020\|EPI_ISL_618032\|2020 | ···················· | ···················· | ····················· | ···················· | ···················· | ···················· |
| P | 4895 | hCoV-19/Denmark/DCGC-8478/2020\|EPI_ISL_618036\|2020 | ···················· | ···················· | ····················· | ···················· | ···················· | ···················· |
| P | 4896 | hCoV-19/Denmark/DCGC-8479/2020\|EPI_ISL_618037\|2020 | ···················· | ···················· | ····················· | ···················· | ···················· | ···················· |
| P | 4897 | hCoV-19/Denmark/DCGC-1262/2020\|EPI_ISL_618039\|2020 | ···················· | ···················· | ····················· | ···················· | ···················· | ···················· |
| P | 4898 | hCoV-19/Denmark/DCGC-7302/2020\|EPI_ISL_619300\|2020 | ···················· | ···················· | ····················· | ···················· | ···················· | ···················· |
| P | 4899 | hCoV-19/Denmark/DCGC-7310/2020\|EPI_ISL_619301\|2020 | ···················· | ···················· | ····················· | ···················· | ···················· | ···················· |
| P | 4900 | hCoV-19/Denmark/DCGC-7323/2020\|EPI_ISL_619302\|2020 | ···················· | ···················· | ····················· | ···················· | ···················· | ···················· |
| P | 4901 | hCoV-19/Denmark/DCGC-7336/2020\|EPI_ISL_619303\|2020 | ···················· | ···················· | ····················· | ···················· | ···················· | ···················· |
| P | 4902 | hCoV-19/Denmark/DCGC-7344/2020\|EPI_ISL_619304\|2020 | ···················· | ···················· | ····················· | ···················· | ···················· | ···················· |
| P | 4903 | hCoV-19/Denmark/DCGC-7352/2020\|EPI_ISL_619305\|2020 | ···················· | ···················· | ····················· | ···················· | ···················· | ···················· |
| P | 4904 | hCoV-19/Denmark/DCGC-7364/2020\|EPI_ISL_619306\|2020 | ···················· | ···················· | ····················· | ···················· | ···················· | ···················· |
| P | 4905 | hCoV-19/Denmark/DCGC-7303/2020\|EPI_ISL_619307\|2020 | ···················· | ···················· | ····················· | ···················· | ···················· | ···················· |
| P | 4906 | hCoV-19/Denmark/DCGC-7337/2020\|EPI_ISL_619308\|2020 | ···················· | ···················· | ····················· | ···················· | ···················· | ···················· |
| P | 4907 | hCoV-19/Denmark/DCGC-7365/2020\|EPI_ISL_619309\|2020 | ···················· | ···················· | ····················· | ···················· | ···················· | ···················· |
| P | 4908 | hCoV-19/Denmark/DCGC-7427/2020\|EPI_ISL_619330\|2020 | ···················· | ···················· | ····················· | ···················· | ···················· | ···················· |
| P | 4909 | hCoV-19/Denmark/DCGC-7447/2020\|EPI_ISL_619331\|2020 | ···················· | ···················· | ····················· | ···················· | ···················· | ···················· |
| P | 4910 | hCoV-19/Denmark/DCGC-1612/2020\|EPI_ISL_619332\|2020 | ···················· | ···················· | ····················· | ···················· | ···················· | ···················· |
| P | 4911 | hCoV-19/Denmark/DCGC-7393/2020\|EPI_ISL_619333\|2020 | ···················· | ···················· | ····················· | ···················· | ···················· | ···················· |
| P | 4912 | hCoV-19/Denmark/DCGC-7413/2020\|EPI_ISL_619334\|2020 | ···················· | ···················· | ····················· | ···················· | ···················· | ···················· |
| P | 4913 | hCoV-19/Denmark/DCGC-7428/2020\|EPI_ISL_619335\|2020 | ···················· | ···················· | ····················· | ···················· | ···················· | ···················· |
| P | 4914 | hCoV-19/Denmark/DCGC-7383/2020\|EPI_ISL_619336\|2020 | ···················· | ···················· | ····················· | ···················· | ···················· | ···················· |
| P | 4915 | hCoV-19/Denmark/DCGC-2046/2020\|EPI_ISL_619337\|2020 | ···················· | ···················· | ····················· | ···················· | ···················· | ···················· |
| P | 4916 | hCoV-19/Denmark/DCGC-7394/2020\|EPI_ISL_619338\|2020 | ···················· | ···················· | ····················· | ···················· | ···················· | ···················· |
| P | 4917 | hCoV-19/Denmark/DCGC-7400/2020\|EPI_ISL_619339\|2020 | ···················· | ···················· | ····················· | ···················· | ···················· | ···················· |

| | | | | | | | | |
|---|---|---|---|---|---|---|---|---|
| P | 4918 | hCoV-19/Denmark/DCGC-7414/2020\|EPI_ISL_619340\|2020 | ···················· | ···················· | ····················· | ···················· | ···················· | ···················· |
| P | 4919 | hCoV-19/Denmark/DCGC-7429/2020\|EPI_ISL_619341\|2020 | ···················· | ···················· | ····················· | ···················· | ···················· | ···················· |
| P | 4920 | hCoV-19/Denmark/DCGC-7441/2020\|EPI_ISL_619342\|2020 | ···················· | ···················· | ····················· | ···················· | ···················· | ···················· |
| P | 4921 | hCoV-19/Denmark/DCGC-7448/2020\|EPI_ISL_619343\|2020 | ···················· | ···················· | ····················· | ···················· | ···················· | ···················· |
| P | 4922 | hCoV-19/Denmark/DCGC-1611/2020\|EPI_ISL_619344\|2020 | ···················· | ···················· | ····················· | ···················· | ···················· | ···················· |
| P | 4923 | hCoV-19/Denmark/DCGC-7384/2020\|EPI_ISL_619345\|2020 | ···················· | ···················· | ····················· | ···················· | ···················· | ···················· |
| P | 4924 | hCoV-19/Denmark/DCGC-7422/2020\|EPI_ISL_619346\|2020 | ···················· | ···················· | ····················· | ···················· | ···················· | ···················· |
| P | 4925 | hCoV-19/Denmark/DCGC-7430/2020\|EPI_ISL_619347\|2020 | ···················· | ···················· | ····················· | ···················· | ···················· | ···················· |
| P | 4926 | hCoV-19/Denmark/DCGC-7442/2020\|EPI_ISL_619348\|2020 | ···················· | ···················· | ····················· | ···················· | ···················· | ···················· |
| P | 4927 | hCoV-19/Denmark/DCGC-7385/2020\|EPI_ISL_619349\|2020 | ···················· | ···················· | ····················· | ···················· | ···················· | ···················· |
| P | 4928 | hCoV-19/Denmark/DCGC-7389/2020\|EPI_ISL_619350\|2020 | ···················· | ···················· | ····················· | ···················· | ···················· | ···················· |
| P | 4929 | hCoV-19/Denmark/DCGC-7396/2020\|EPI_ISL_619351\|2020 | ···················· | ···················· | ····················· | ···················· | ···················· | ···················· |
| P | 4930 | hCoV-19/Denmark/DCGC-7423/2020\|EPI_ISL_619352\|2020 | ···················· | ···················· | ····················· | ···················· | ···················· | ···················· |
| P | 4931 | hCoV-19/Denmark/DCGC-7431/2020\|EPI_ISL_619353\|2020 | ···················· | ····A··············· | ····················· | ···················· | ···················· | ···················· |
| P | 4932 | hCoV-19/Denmark/DCGC-7443/2020\|EPI_ISL_619354\|2020 | ···················· | ···················· | ····················· | ···················· | ···················· | ···················· |
| P | 4933 | hCoV-19/Denmark/DCGC-7449/2020\|EPI_ISL_619355\|2020 | ···················· | ···················· | ····················· | ···················· | ···················· | ···················· |
| P | 4934 | hCoV-19/Denmark/DCGC-7390/2020\|EPI_ISL_619356\|2020 | ···················· | ···················· | ····················· | ···················· | ···················· | ···················· |
| P | 4935 | hCoV-19/Denmark/DCGC-7397/2020\|EPI_ISL_619357\|2020 | ···················· | ···················· | ····················· | ···················· | ···················· | ···················· |
| P | 4936 | hCoV-19/Denmark/DCGC-7402/2020\|EPI_ISL_619358\|2020 | ···················· | ·A·················· | ····················· | ···················· | ···················· | ···················· |
| P | 4937 | hCoV-19/Denmark/DCGC-7409/2020\|EPI_ISL_619359\|2020 | ···················· | ···················· | ····················· | ···················· | ···················· | ···················· |
| P | 4938 | hCoV-19/Denmark/DCGC-7417/2020\|EPI_ISL_619360\|2020 | ···················· | ···················· | ····················· | ···················· | ···················· | ···················· |
| P | 4939 | hCoV-19/Denmark/DCGC-7432/2020\|EPI_ISL_619361\|2020 | ···················· | ···················· | ····················· | ···················· | ···················· | ···················· |
| P | 4940 | hCoV-19/Denmark/DCGC-7444/2020\|EPI_ISL_619362\|2020 | ···················· | ···················· | ····················· | ···················· | ···················· | ···················· |
| P | 4941 | hCoV-19/Denmark/DCGC-7391/2020\|EPI_ISL_619363\|2020 | ···················· | ···················· | ····················· | ···················· | ···················· | ···················· |
| P | 4942 | hCoV-19/Denmark/DCGC-7410/2020\|EPI_ISL_619364\|2020 | ···················· | ·A·················· | ····················· | ···················· | ···················· | ···················· |
| P | 4943 | hCoV-19/Denmark/DCGC-7418/2020\|EPI_ISL_619365\|2020 | ···················· | ···················· | ····················· | ···················· | ···················· | ···················· |
| P | 4944 | hCoV-19/Denmark/DCGC-7425/2020\|EPI_ISL_619366\|2020 | ···················· | ···················· | ····················· | ···················· | ···················· | ···················· |
| P | 4945 | hCoV-19/Denmark/DCGC-7433/2020\|EPI_ISL_619367\|2020 | ···················· | ·A·················· | ····················· | ···················· | ···················· | ···················· |
| P | 4946 | hCoV-19/Denmark/DCGC-7445/2020\|EPI_ISL_619368\|2020 | ···················· | ···················· | ····················· | ···················· | ···················· | ···················· |
| P | 4947 | hCoV-19/Denmark/DCGC-7455/2020\|EPI_ISL_619369\|2020 | ···················· | ···················· | ····················· | ···················· | ···················· | ···················· |
| P | 4948 | hCoV-19/Denmark/DCGC-7459/2020\|EPI_ISL_619400\|2020 | ···················· | ···················· | ····················· | ···················· | ···················· | ···················· |
| P | 4949 | hCoV-19/Denmark/DCGC-7466/2020\|EPI_ISL_619401\|2020 | ···················· | ···················· | ····················· | ···················· | ···················· | ···················· |
| P | 4950 | hCoV-19/Denmark/DCGC-7472/2020\|EPI_ISL_619402\|2020 | ···················· | ···················· | ····················· | ···················· | ···················· | ···················· |
| P | 4951 | hCoV-19/Denmark/DCGC-7482/2020\|EPI_ISL_619403\|2020 | ···················· | ···················· | ····················· | ···················· | ···················· | ···················· |
| P | 4952 | hCoV-19/Denmark/DCGC-7490/2020\|EPI_ISL_619404\|2020 | ···················· | ···················· | ····················· | ···················· | ···················· | ···················· |
| P | 4953 | hCoV-19/Denmark/DCGC-1607/2020\|EPI_ISL_619405\|2020 | ···················· | ···················· | ····················· | ···················· | ···················· | ···················· |
| P | 4954 | hCoV-19/Denmark/DCGC-6506/2020\|EPI_ISL_619406\|2020 | ···················· | ···················· | ····················· | ···················· | ···················· | ···················· |
| P | 4955 | hCoV-19/Denmark/DCGC-6507/2020\|EPI_ISL_619407\|2020 | ···················· | ···················· | ····················· | ···················· | ···················· | ···················· |
| P | 4956 | hCoV-19/Denmark/DCGC-6508/2020\|EPI_ISL_619408\|2020 | ···················· | ···················· | ····················· | ···················· | ···················· | ···················· |
| P | 4957 | hCoV-19/Denmark/DCGC-6509/2020\|EPI_ISL_619409\|2020 | ···················· | ···················· | ····················· | ···················· | ···················· | ···················· |
| P | 4958 | hCoV-19/Denmark/DCGC-6408/2020\|EPI_ISL_620308\|2020 | ···················· | ·A·················· | ····················· | ···················· | ···················· | ···················· |
| P | 4959 | hCoV-19/Denmark/DCGC-6409/2020\|EPI_ISL_620309\|2020 | ···················· | ···················· | ····················· | ···················· | ···················· | ···················· |
| P | 4960 | hCoV-19/Denmark/DCGC-6414/2020\|EPI_ISL_620314\|2020 | ···················· | ···················· | ····················· | ···················· | ···················· | ···················· |
| P | 4961 | hCoV-19/Denmark/DCGC-6415/2020\|EPI_ISL_620315\|2020 | ···················· | ···················· | ····················· | ···················· | ···················· | ···················· |
| P | 4962 | hCoV-19/Denmark/DCGC-6416/2020\|EPI_ISL_620316\|2020 | ···················· | ···················· | ····················· | ···················· | ···················· | ···················· |
| P | 4963 | hCoV-19/Denmark/DCGC-6417/2020\|EPI_ISL_620317\|2020 | ···················· | ···················· | ····················· | ···················· | ···················· | ···················· |
| P | 4964 | hCoV-19/Denmark/DCGC-6418/2020\|EPI_ISL_620318\|2020 | ···················· | ···················· | ····················· | ···················· | ···················· | ···················· |
| P | 4965 | hCoV-19/Denmark/DCGC-6419/2020\|EPI_ISL_620319\|2020 | ···················· | ···················· | ····················· | ···················· | ···················· | ···················· |
| P | 4966 | hCoV-19/Denmark/DCGC-6420/2020\|EPI_ISL_620320\|2020 | ···················· | ···················· | ····················· | ···················· | ···················· | ···················· |
| P | 4967 | hCoV-19/Denmark/DCGC-6421/2020\|EPI_ISL_620321\|2020 | ···················· | ···················· | ····················· | ···················· | ···················· | ···················· |
| P | 4968 | hCoV-19/Denmark/DCGC-6422/2020\|EPI_ISL_620322\|2020 | ···················· | ···················· | ····················· | ···················· | ···················· | ···················· |
| P | 4969 | hCoV-19/Denmark/DCGC-6423/2020\|EPI_ISL_620323\|2020 | ···················· | ···················· | ····················· | ···················· | ···················· | ···················· |
| P | 4970 | hCoV-19/Denmark/DCGC-6424/2020\|EPI_ISL_620324\|2020 | ···················· | ···················· | ····················· | ···················· | ···················· | ···················· |
| P | 4971 | hCoV-19/Denmark/DCGC-6425/2020\|EPI_ISL_620325\|2020 | ···················· | ···················· | ····················· | ···················· | ···················· | ···················· |
| P | 4972 | hCoV-19/Denmark/DCGC-6426/2020\|EPI_ISL_620326\|2020 | ···················· | ···················· | ····················· | ···················· | ···················· | ···················· |
| P | 4973 | hCoV-19/Denmark/DCGC-6428/2020\|EPI_ISL_620328\|2020 | ···················· | ···················· | ····················· | ···················· | ···················· | ···················· |
| P | 4974 | hCoV-19/Denmark/DCGC-6429/2020\|EPI_ISL_620329\|2020 | ···················· | ···················· | ····················· | ···················· | ···················· | ···················· |
| P | 4975 | hCoV-19/Denmark/DCGC-6430/2020\|EPI_ISL_620330\|2020 | ···················· | ···················· | ····················· | ···················· | ···················· | ···················· |
| P | 4976 | hCoV-19/Denmark/DCGC-6431/2020\|EPI_ISL_620331\|2020 | ···················· | ···················· | ····················· | ···················· | ···················· | ···················· |
| P | 4977 | hCoV-19/Denmark/DCGC-6433/2020\|EPI_ISL_620333\|2020 | ···················· | ···················· | ····················· | ···················· | ···················· | ···················· |
| P | 4978 | hCoV-19/Denmark/DCGC-6435/2020\|EPI_ISL_620335\|2020 | ···················· | ···················· | ····················· | ···················· | ···················· | ···················· |
| P | 4979 | hCoV-19/Denmark/DCGC-6436/2020\|EPI_ISL_620336\|2020 | ···················· | ···················· | ····················· | ···················· | ···················· | ···················· |
| P | 4980 | hCoV-19/Denmark/DCGC-6437/2020\|EPI_ISL_620337\|2020 | ···················· | ···················· | ····················· | ···················· | ···················· | ···················· |
| P | 4981 | hCoV-19/Denmark/DCGC-6438/2020\|EPI_ISL_620338\|2020 | ···················· | ···················· | ····················· | ···················· | ···················· | ···················· |
| P | 4982 | hCoV-19/Denmark/DCGC-6439/2020\|EPI_ISL_620339\|2020 | ···················· | ···················· | ····················· | ···················· | ···················· | ···················· |
| P | 4983 | hCoV-19/Denmark/DCGC-6440/2020\|EPI_ISL_620340\|2020 | ···················· | ···················· | ····················· | ···················· | ···················· | ···················· |
| P | 4984 | hCoV-19/Denmark/DCGC-6441/2020\|EPI_ISL_620341\|2020 | ···················· | ···················· | ····················· | ···················· | ······T············· | ···················· |
| P | 4985 | hCoV-19/Denmark/DCGC-6442/2020\|EPI_ISL_620342\|2020 | ···················· | ···················· | ····················· | ···················· | ···················· | ···················· |
| P | 4986 | hCoV-19/Denmark/DCGC-6443/2020\|EPI_ISL_620343\|2020 | ···················· | ···················· | ····················· | ···················· | ···················· | ···················· |
| P | 4987 | hCoV-19/Denmark/DCGC-6444/2020\|EPI_ISL_620344\|2020 | ···················· | ···················· | ····················· | ···················· | ···················· | ···················· |
| P | 4988 | hCoV-19/Denmark/DCGC-6448/2020\|EPI_ISL_620348\|2020 | ···················· | ···················· | ····················· | ···················· | ···················· | ···················· |
| P | 4989 | hCoV-19/Denmark/DCGC-6450/2020\|EPI_ISL_620350\|2020 | ···················· | ···················· | ····················· | ···················· | ···················· | ···················· |
| P | 4990 | hCoV-19/Denmark/DCGC-6451/2020\|EPI_ISL_620351\|2020 | ···················· | ···················· | ····················· | ···················· | ···················· | ···················· |
| P | 4991 | hCoV-19/Denmark/DCGC-6452/2020\|EPI_ISL_620352\|2020 | ···················· | ···················· | ····················· | ···················· | ···················· | ···················· |
| P | 4992 | hCoV-19/Denmark/DCGC-6454/2020\|EPI_ISL_620354\|2020 | ···················· | ···················· | ····················· | ···················· | ···················· | ···················· |
| P | 4993 | hCoV-19/Denmark/DCGC-6455/2020\|EPI_ISL_620355\|2020 | ···················· | ···················· | ····················· | ···················· | ···················· | ···················· |
| P | 4994 | hCoV-19/Denmark/DCGC-6456/2020\|EPI_ISL_620356\|2020 | ···················· | ···················· | ····················· | ···················· | ···················· | ···················· |
| P | 4995 | hCoV-19/Denmark/DCGC-6457/2020\|EPI_ISL_620357\|2020 | ···················· | ···················· | ····················· | ···················· | ···················· | ···················· |
| P | 4996 | hCoV-19/Denmark/DCGC-5063/2020\|EPI_ISL_621601\|2020 | ···················· | ···················· | ····················· | ···················· | ···················· | ···················· |
| P | 4997 | hCoV-19/Denmark/DCGC-5064/2020\|EPI_ISL_621602\|2020 | ···················· | ···················· | ····················· | ···················· | ···················· | ···················· |

| | | | | | | | | |
|---|---|---|---|---|---|---|---|---|
| P | 4998 | hCoV-19/Denmark/DCGC-5065/2020\|EPI_ISL_621603\|2020 | ···················· | ···················· | ···················· | ···················· | ···················· | ···················· |
| P | 4999 | hCoV-19/Denmark/DCGC-5066/2020\|EPI_ISL_621604\|2020 | ···················· | ···················· | ···················· | ···················· | ···················· | ···················· |
| P | 5000 | hCoV-19/Denmark/DCGC-5067/2020\|EPI_ISL_621605\|2020 | ···················· | ···················· | ···················· | ···················· | ···················· | ···················· |
| P | 5001 | hCoV-19/Denmark/DCGC-5070/2020\|EPI_ISL_621608\|2020 | ···················· | ···················· | ···················· | ···················· | ···················· | ···················· |
| P | 5002 | hCoV-19/Denmark/DCGC-5071/2020\|EPI_ISL_621609\|2020 | ···················· | ···················· | ···················· | ···················· | ···················· | ···················· |
| P | 5003 | hCoV-19/Denmark/DCGC-5072/2020\|EPI_ISL_621610\|2020 | ···················· | ···················· | ···················· | ···················· | ···················· | ···················· |
| P | 5004 | hCoV-19/Denmark/DCGC-5075/2020\|EPI_ISL_621613\|2020 | ···················· | ···················· | ···················· | ···················· | ···················· | ···················· |
| P | 5005 | hCoV-19/Denmark/DCGC-5076/2020\|EPI_ISL_621614\|2020 | ···················· | ···················· | ···················· | ···················· | ···················· | ···················· |
| P | 5006 | hCoV-19/Denmark/DCGC-5078/2020\|EPI_ISL_621616\|2020 | ···················· | ···················· | ···················· | ···················· | ···················· | ···················· |
| P | 5007 | hCoV-19/Denmark/DCGC-5079/2020\|EPI_ISL_621617\|2020 | ···················· | ···················· | ···················· | ···················· | ···················· | ···················· |
| P | 5008 | hCoV-19/Denmark/DCGC-5080/2020\|EPI_ISL_621618\|2020 | ···················· | ···················· | ···················· | ···················· | ···················· | ···················· |
| P | 5009 | hCoV-19/Denmark/DCGC-5081/2020\|EPI_ISL_621619\|2020 | ···················· | ···················· | ···················· | ···················· | ···················· | ···················· |
| P | 5010 | hCoV-19/Denmark/DCGC-5082/2020\|EPI_ISL_621620\|2020 | ···················· | ···················· | ···················· | ···················· | ···················· | ···················· |
| P | 5011 | hCoV-19/Denmark/DCGC-5083/2020\|EPI_ISL_621621\|2020 | ···················· | ···················· | ···················· | ···················· | ···················· | ···················· |
| P | 5012 | hCoV-19/Denmark/DCGC-5101/2020\|EPI_ISL_621639\|2020 | ···················· | ···················· | ···················· | ···················· | ···················· | ···················· |
| P | 5013 | hCoV-19/Denmark/DCGC-5107/2020\|EPI_ISL_621645\|2020 | ···················· | ···················· | ···················· | ···················· | ···················· | ···················· |
| P | 5014 | hCoV-19/Denmark/DCGC-5108/2020\|EPI_ISL_621646\|2020 | ···················· | ···················· | ···················· | ···················· | ···················· | ···················· |
| P | 5015 | hCoV-19/Denmark/DCGC-5110/2020\|EPI_ISL_621648\|2020 | ···················· | ···················· | ···················· | ···················· | ···················· | ···················· |
| P | 5016 | hCoV-19/Denmark/DCGC-5111/2020\|EPI_ISL_621649\|2020 | ···················· | ···················· | ···················· | ···················· | ···················· | ···················· |
| P | 5017 | hCoV-19/Denmark/DCGC-5112/2020\|EPI_ISL_621650\|2020 | ···················· | ···················· | ···················· | ···················· | ···················· | ···················· |
| P | 5018 | hCoV-19/Denmark/DCGC-5113/2020\|EPI_ISL_621651\|2020 | ···················· | ···················· | ···················· | ···················· | ···················· | ···················· |
| P | 5019 | hCoV-19/Denmark/DCGC-5114/2020\|EPI_ISL_621652\|2020 | ···················· | ···················· | ···················· | ···················· | ···················· | ···················· |
| P | 5020 | hCoV-19/Denmark/DCGC-5115/2020\|EPI_ISL_621653\|2020 | ···················· | ···················· | ···················· | ···················· | ···················· | ···················· |
| P | 5021 | hCoV-19/Denmark/DCGC-5116/2020\|EPI_ISL_621654\|2020 | ···················· | ···················· | ···················· | ···················· | ···················· | ···················· |
| P | 5022 | hCoV-19/Denmark/DCGC-5117/2020\|EPI_ISL_621655\|2020 | ···················· | ···················· | ···················· | ···················· | ···················· | ···················· |
| P | 5023 | hCoV-19/Denmark/DCGC-5118/2020\|EPI_ISL_621656\|2020 | ···················· | ···················· | ···················· | ···················· | ···················· | ···················· |
| P | 5024 | hCoV-19/Denmark/DCGC-5119/2020\|EPI_ISL_621657\|2020 | ···················· | ···················· | ···················· | ···················· | ···················· | ···················· |
| P | 5025 | hCoV-19/Denmark/DCGC-5120/2020\|EPI_ISL_621658\|2020 | ···················· | ···················· | ···················· | ···················· | ···················· | ···················· |
| P | 5026 | hCoV-19/Denmark/DCGC-5121/2020\|EPI_ISL_621659\|2020 | ···················· | ···················· | ···················· | ···················· | ···················· | ···················· |
| P | 5027 | hCoV-19/Denmark/DCGC-5123/2020\|EPI_ISL_621661\|2020 | ···················· | ·A·················· | ···················· | ···················· | ···················· | ···················· |
| P | 5028 | hCoV-19/Denmark/DCGC-5124/2020\|EPI_ISL_621662\|2020 | ···················· | ···················· | ···················· | ···················· | ···················· | ···················· |
| P | 5029 | hCoV-19/Denmark/DCGC-5126/2020\|EPI_ISL_621664\|2020 | ···················· | ···················· | ···················· | ···················· | ···················· | ···················· |
| P | 5030 | hCoV-19/Denmark/DCGC-5127/2020\|EPI_ISL_621665\|2020 | ···················· | ···················· | ···················· | ···················· | ···················· | ···················· |
| P | 5031 | hCoV-19/Denmark/DCGC-5129/2020\|EPI_ISL_621667\|2020 | ···················· | ···················· | ···················· | ···················· | ···················· | ···················· |
| P | 5032 | hCoV-19/Denmark/DCGC-5130/2020\|EPI_ISL_621668\|2020 | ···················· | ···················· | ···················· | ···················· | ···················· | ···················· |
| P | 5033 | hCoV-19/Denmark/DCGC-5131/2020\|EPI_ISL_621669\|2020 | ···················· | ···················· | ···················· | ···················· | ···················· | ···················· |
| P | 5034 | hCoV-19/Denmark/DCGC-5132/2020\|EPI_ISL_621670\|2020 | ···················· | ···················· | ···················· | ···················· | ···················· | ···················· |
| P | 5035 | hCoV-19/Denmark/DCGC-5133/2020\|EPI_ISL_621671\|2020 | ···················· | ···················· | ···················· | ···················· | ···················· | ···················· |
| P | 5036 | hCoV-19/Denmark/DCGC-5134/2020\|EPI_ISL_621672\|2020 | ···················· | ···················· | ···················· | ···················· | ···················· | ···················· |
| P | 5037 | hCoV-19/Denmark/DCGC-5135/2020\|EPI_ISL_621673\|2020 | ···················· | ···················· | ···················· | ···················· | ···················· | ···················· |
| P | 5038 | hCoV-19/Denmark/DCGC-5136/2020\|EPI_ISL_621674\|2020 | ···················· | ···················· | ···················· | ···················· | ···················· | ···················· |
| P | 5039 | hCoV-19/Denmark/DCGC-5137/2020\|EPI_ISL_621675\|2020 | ···················· | ···················· | ···················· | ···················· | ···················· | ···················· |
| P | 5040 | hCoV-19/Denmark/DCGC-5139/2020\|EPI_ISL_621677\|2020 | ···················· | ···················· | ···················· | ···················· | ···················· | ···················· |
| P | 5041 | hCoV-19/Denmark/DCGC-5140/2020\|EPI_ISL_621678\|2020 | ···················· | ···················· | ···················· | ···················· | ···················· | ···················· |
| P | 5042 | hCoV-19/Denmark/DCGC-5141/2020\|EPI_ISL_621679\|2020 | ···················· | ···················· | ···················· | ···················· | ···················· | ···················· |
| P | 5043 | hCoV-19/Denmark/DCGC-5142/2020\|EPI_ISL_621680\|2020 | ···················· | ···················· | ···················· | ···················· | ···················· | ···················· |
| P | 5044 | hCoV-19/Denmark/DCGC-5143/2020\|EPI_ISL_621681\|2020 | ···················· | ···················· | ···················· | ···················· | ···················· | ···················· |
| P | 5045 | hCoV-19/Denmark/DCGC-5144/2020\|EPI_ISL_621682\|2020 | ···················· | ···················· | ···················· | ···················· | ···················· | ···················· |
| P | 5046 | hCoV-19/Denmark/DCGC-5145/2020\|EPI_ISL_621683\|2020 | ···················· | ···················· | ···················· | ···················· | ···················· | ···················· |
| P | 5047 | hCoV-19/Denmark/DCGC-5146/2020\|EPI_ISL_621684\|2020 | ···················· | ···················· | ···················· | ···················· | ···················· | ···················· |
| P | 5048 | hCoV-19/Denmark/DCGC-5148/2020\|EPI_ISL_621686\|2020 | ···················· | ···················· | ···················· | ···················· | ···················· | ···················· |
| P | 5049 | hCoV-19/Denmark/DCGC-5149/2020\|EPI_ISL_621687\|2020 | ···················· | ···················· | ···················· | ···················· | ···················· | ···················· |
| P | 5050 | hCoV-19/Denmark/DCGC-5150/2020\|EPI_ISL_621688\|2020 | ···················· | ···················· | ···················· | ···················· | ···················· | ···················· |
| P | 5051 | hCoV-19/Denmark/DCGC-5162/2020\|EPI_ISL_621700\|2020 | ···················· | ···················· | ···················· | ···················· | ···················· | ···················· |
| P | 5052 | hCoV-19/Denmark/DCGC-5163/2020\|EPI_ISL_621701\|2020 | ···················· | ···················· | ···················· | ···················· | ···················· | ···················· |
| P | 5053 | hCoV-19/Denmark/DCGC-5164/2020\|EPI_ISL_621702\|2020 | ···················· | ···················· | ···················· | ···················· | ···················· | ···················· |
| P | 5054 | hCoV-19/Denmark/DCGC-5165/2020\|EPI_ISL_621703\|2020 | ···················· | ···················· | ···················· | ···················· | ···················· | ···················· |
| P | 5055 | hCoV-19/Denmark/DCGC-5166/2020\|EPI_ISL_621704\|2020 | ···················· | ···················· | ···················· | ···················· | ···················· | ···················· |
| P | 5056 | hCoV-19/Denmark/DCGC-5167/2020\|EPI_ISL_621705\|2020 | ···················· | ···················· | ···················· | ···················· | ···················· | ···················· |
| P | 5057 | hCoV-19/Denmark/DCGC-5168/2020\|EPI_ISL_621706\|2020 | ···················· | ···················· | ···················· | ···················· | ···················· | ···················· |
| P | 5058 | hCoV-19/Denmark/DCGC-5169/2020\|EPI_ISL_621707\|2020 | ···················· | ·A·················· | ···················· | ···················· | ···················· | ···················· |
| P | 5059 | hCoV-19/Denmark/DCGC-5171/2020\|EPI_ISL_621709\|2020 | ···················· | ···················· | ···················· | ···················· | ···················· | ···················· |
| P | 5060 | hCoV-19/Denmark/DCGC-5172/2020\|EPI_ISL_621710\|2020 | ···················· | ·A·················· | ···················· | ···················· | ···················· | ···················· |
| P | 5061 | hCoV-19/Denmark/DCGC-5173/2020\|EPI_ISL_621711\|2020 | ···················· | ···················· | ···················· | ···················· | ···················· | ···················· |
| P | 5062 | hCoV-19/Denmark/DCGC-5174/2020\|EPI_ISL_621712\|2020 | ···················· | ···················· | ···················· | ···················· | ···················· | ···················· |
| P | 5063 | hCoV-19/Denmark/DCGC-5175/2020\|EPI_ISL_621713\|2020 | ···················· | ···················· | ···················· | ···················· | ···················· | ···················· |
| P | 5064 | hCoV-19/Denmark/DCGC-5176/2020\|EPI_ISL_621714\|2020 | ···················· | ···················· | ···················· | ···················· | ···················· | ···················· |
| P | 5065 | hCoV-19/Denmark/DCGC-5177/2020\|EPI_ISL_621715\|2020 | ···················· | ···················· | ···················· | ···················· | ···················· | ···················· |
| P | 5066 | hCoV-19/Denmark/DCGC-5181/2020\|EPI_ISL_621719\|2020 | ········T··········· | ···················· | ···················· | ···················· | ···················· | ···················· |
| P | 5067 | hCoV-19/Denmark/DCGC-5182/2020\|EPI_ISL_621720\|2020 | ···················· | ···················· | ···················· | ···················· | ···················· | ···················· |
| P | 5068 | hCoV-19/Denmark/DCGC-5183/2020\|EPI_ISL_621721\|2020 | ···················· | ···················· | ···················· | ···················· | ···················· | ···················· |
| P | 5069 | hCoV-19/South Africa/R13067-20/2020\|EPI_ISL_622934 | ···················· | ·A·················· | ···················· | ···················· | ···················· | ···················· |
| P | 5070 | hCoV-19/South Africa/R13147-20/2020\|EPI_ISL_622935 | ···················· | ·A·················· | ···················· | ···················· | ···················· | ···················· |
| P | 5071 | hCoV-19/South Africa/R13159-20/2020\|EPI_ISL_622936 | ···················· | ···················· | ···················· | ···················· | ···················· | ···················· |
| P | 5072 | hCoV-19/South Africa/R13274-20/2020\|EPI_ISL_622937 | ···················· | ···················· | ···················· | ···················· | ···················· | ···················· |
| P | 5073 | hCoV-19/South Africa/R13278-20/2020\|EPI_ISL_622938 | ···················· | ···················· | ···················· | ···················· | ···················· | ···················· |
| P | 5074 | hCoV-19/South Africa/R13317-20/2020\|EPI_ISL_622939 | ···················· | ···················· | ···················· | ···················· | ···················· | ···················· |
| P | 5075 | hCoV-19/South Africa/R13452-20/2020\|EPI_ISL_622940 | ···················· | ···················· | ···················· | ···················· | ···················· | ···················· |
| P | 5076 | hCoV-19/South Africa/R13455-20/2020\|EPI_ISL_622941 | ···················· | ···················· | ···················· | ···················· | ···················· | ···················· |
| P | 5077 | hCoV-19/England/ALDP-AA2D3C/2020\|EPI_ISL_625250\|20 | ···················· | ···················· | ···················· | ···················· | ···················· | ···················· |

| | | | | | | | |
|---|---|---|---|---|---|---|---|
| P | 5078 hCoV-19/England/ALDP-AA1896/2020\|EPI_ISL_625251\|20 | ···················· | ···················· | ···················· | ···················· | ···················· | ···················· |
| P | 5079 hCoV-19/England/ALDP-AA558C/2020\|EPI_ISL_625252\|20 | ···················· | ···················· | ···················· | ···················· | ···················· | ···················· |
| P | 5080 hCoV-19/England/ALDP-AA35CA/2020\|EPI_ISL_625253\|20 | ···················· | ·A·················· | ···················· | ···················· | ···················· | ···A················ |
| P | 5081 hCoV-19/England/ALDP-AA1E58/2020\|EPI_ISL_625254\|20 | ···················· | ···················· | ···················· | ···················· | ···················· | ···················· |
| P | 5082 hCoV-19/England/ALDP-AA530A/2020\|EPI_ISL_625255\|20 | ···················· | ······A············· | ···················· | ···················· | ···················· | ···················· |
| P | 5083 hCoV-19/England/ALDP-AA5522/2020\|EPI_ISL_625257\|20 | ···················· | ···················· | ···················· | ···················· | ···················· | ···················· |
| P | 5084 hCoV-19/England/ALDP-AA1623/2020\|EPI_ISL_625258\|20 | ···················· | ···················· | ···················· | ···················· | ···················· | ···················· |
| P | 5085 hCoV-19/England/ALDP-AA5355/2020\|EPI_ISL_625259\|20 | ···················· | ·········T·········· | ···················· | ···················· | ···················· | ···················· |
| P | 5086 hCoV-19/England/ALDP-AA458D/2020\|EPI_ISL_625260\|20 | ···················· | ···················· | ···················· | ···················· | ···················· | ···················· |
| P | 5087 hCoV-19/England/ALDP-AA2CC6/2020\|EPI_ISL_625261\|20 | ···········T········ | ···················· | ···················· | ···················· | ···················· | ···················· |
| P | 5088 hCoV-19/England/ALDP-AA21D6/2020\|EPI_ISL_625262\|20 | ···················· | ···················· | ···················· | ···················· | ···················· | ···················· |
| P | 5089 hCoV-19/England/ALDP-AA3296/2020\|EPI_ISL_625263\|20 | ···················· | ···················· | ···················· | ···················· | ···················· | ···················· |
| P | 5090 hCoV-19/England/ALDP-AA2C99/2020\|EPI_ISL_625264\|20 | ···················· | ···················· | ···················· | ···················· | ···················· | ···················· |
| P | 5091 hCoV-19/England/ALDP-AA37E2/2020\|EPI_ISL_625265\|20 | ···················· | ···················· | ···················· | ···················· | ···················· | ···················· |
| P | 5092 hCoV-19/England/ALDP-AA4A33/2020\|EPI_ISL_625266\|20 | ···················· | ···················· | ···················· | ···················· | ···················· | ···················· |
| P | 5093 hCoV-19/Wales/ALDP-A851E8/2020\|EPI_ISL_625267\|2020 | ···················· | ···················· | ···················· | ···················· | ···················· | ···················· |
| P | 5094 hCoV-19/Wales/ALDP-A85160/2020\|EPI_ISL_625268\|2020 | ···················· | ···················· | ···················· | ···················· | ···················· | ···················· |
| P | 5095 hCoV-19/Wales/ALDP-A8517F/2020\|EPI_ISL_625269\|2020 | ···················· | ···················· | ···················· | ···················· | ···················· | ···················· |
| P | 5096 hCoV-19/Wales/ALDP-A8519D/2020\|EPI_ISL_625270\|2020 | ···················· | ···················· | ···················· | ···················· | ···················· | ···················· |
| P | 5097 hCoV-19/Wales/ALDP-A85810/2020\|EPI_ISL_625271\|2020 | ···················· | ···················· | ···················· | ···················· | ···················· | ···················· |
| P | 5098 hCoV-19/Wales/ALDP-A85115/2020\|EPI_ISL_625272\|2020 | ···················· | ···················· | ···················· | ···················· | ···················· | ···················· |
| P | 5099 hCoV-19/Wales/ALDP-A851D9/2020\|EPI_ISL_625273\|2020 | ···················· | ···················· | ···················· | ···················· | ···················· | ···················· |
| P | 5100 hCoV-19/Wales/ALDP-A85133/2020\|EPI_ISL_625274\|2020 | ···················· | ···················· | ···················· | ···················· | ···················· | ···················· |
| P | 5101 hCoV-19/Wales/ALDP-A851F7/2020\|EPI_ISL_625275\|2020 | ···················· | ···················· | ···················· | ···················· | ···················· | ···················· |
| P | 5102 hCoV-19/Wales/ALDP-A85106/2020\|EPI_ISL_625276\|2020 | ···················· | ···················· | ···················· | ···················· | ···················· | ···················· |
| P | 5103 hCoV-19/Wales/ALDP-A85643/2020\|EPI_ISL_625277\|2020 | ···················· | ···················· | ···················· | ···················· | ···················· | ···················· |
| P | 5104 hCoV-19/Wales/ALDP-A85151/2020\|EPI_ISL_625279\|2020 | ···················· | ···················· | ···················· | ···················· | ···················· | ···················· |
| P | 5105 hCoV-19/Wales/ALDP-A85616/2020\|EPI_ISL_625280\|2020 | ···················· | ···················· | ···················· | ···················· | ···················· | ···················· |
| P | 5106 hCoV-19/Scotland/CAMC-A822CA/2020\|EPI_ISL_625281\|2 | ···················· | ···················· | ···················· | ···················· | ···················· | ···················· |
| P | 5107 hCoV-19/Wales/CAMC-A81FA6/2020\|EPI_ISL_625282\|2020 | ···················· | ···················· | ···················· | ···················· | ···················· | ···················· |
| P | 5108 hCoV-19/Scotland/CAMC-A82E20/2020\|EPI_ISL_625283\|2 | ···················· | ···················· | ···················· | ···················· | ···················· | ···················· |
| P | 5109 hCoV-19/Scotland/CAMC-A8237C/2020\|EPI_ISL_625284\|2 | ···················· | ···················· | ···················· | ···················· | ···················· | ···················· |
| P | 5110 hCoV-19/Scotland/CAMC-A824E2/2020\|EPI_ISL_625285\|2 | ···················· | ···················· | ···················· | ···················· | ···················· | ···················· |
| P | 5111 hCoV-19/Wales/CAMC-A823B8/2020\|EPI_ISL_625286\|2020 | ···················· | ···················· | ···················· | ···················· | ···················· | ···················· |
| P | 5112 hCoV-19/Wales/CAMC-A80E31/2020\|EPI_ISL_625287\|2020 | ···················· | ···················· | ···················· | ···················· | ···················· | ···················· |
| P | 5113 hCoV-19/Wales/CAMC-A82D05/2020\|EPI_ISL_625288\|2020 | ···················· | ···················· | ···················· | ···················· | ···················· | ···················· |
| P | 5114 hCoV-19/Wales/CAMC-A82075/2020\|EPI_ISL_625292\|2020 | ···················· | ···················· | ···················· | ···················· | ···················· | ···················· |
| P | 5115 hCoV-19/Scotland/CAMC-A82ED5/2020\|EPI_ISL_625294\|2 | ···················· | ···················· | ···················· | ···················· | ···················· | ···················· |
| P | 5116 hCoV-19/Wales/CAMC-A80DF8/2020\|EPI_ISL_625295\|2020 | ···················· | ···················· | ···················· | ···················· | ···················· | ···················· |
| P | 5117 hCoV-19/Wales/CAMC-A81D24/2020\|EPI_ISL_625296\|2020 | ···················· | ···················· | ···················· | ···················· | ···················· | ···················· |
| P | 5118 hCoV-19/Wales/CAMC-A8244C/2020\|EPI_ISL_625298\|2020 | ···················· | ···················· | ···················· | ···················· | ···················· | ···················· |
| P | 5119 hCoV-19/Scotland/CAMC-A82C08/2020\|EPI_ISL_625299\|2 | ···················· | ···················· | ···················· | ···················· | ···················· | ···················· |
| P | 5120 hCoV-19/Philippines/UPMNIH_C1/2020\|EPI_ISL_634976\| | ···················· | ···················· | ···················· | ···················· | ···················· | ···················· |
| P | 5121 hCoV-19/Denmark/DCGC-1042/2020\|EPI_ISL_614440\|2020 | ···················· | ···················· | ···················· | ···················· | ···················· | ···················· |
| P | 5122 hCoV-19/Denmark/DCGC-1212/2020\|EPI_ISL_614441\|2020 | ···················· | ···················· | ···················· | ···················· | ···················· | ···················· |
| P | 5123 hCoV-19/Denmark/DCGC-1214/2020\|EPI_ISL_614442\|2020 | ···················· | ···················· | ···················· | ···················· | ···················· | ···················· |
| P | 5124 hCoV-19/Denmark/DCGC-786/2020\|EPI_ISL_614443\|2020- | ···················· | ···················· | ···················· | ···················· | ···················· | ···················· |
| P | 5125 hCoV-19/Denmark/DCGC-791/2020\|EPI_ISL_614444\|2020- | ···················· | ···················· | ···················· | ···················· | ···················· | ···················· |
| P | 5126 hCoV-19/Denmark/DCGC-1008/2020\|EPI_ISL_614448\|2020 | ···················· | ···················· | ···················· | ···················· | ···················· | ···················· |
| P | 5127 hCoV-19/Denmark/DCGC-914/2020\|EPI_ISL_614449\|2020- | ···················· | ···················· | ···················· | ···················· | ···················· | ···················· |
| P | 5128 hCoV-19/Denmark/DCGC-1075/2020\|EPI_ISL_614500\|2020 | ···················· | ···················· | ···················· | ···················· | ···················· | ···················· |
| P | 5129 hCoV-19/Denmark/DCGC-1076/2020\|EPI_ISL_614501\|2020 | ···················· | ···················· | ···················· | ···················· | ···················· | ···················· |
| P | 5130 hCoV-19/Denmark/DCGC-1077/2020\|EPI_ISL_614502\|2020 | ···················· | ···················· | ···················· | ···················· | ···················· | ···················· |
| P | 5131 hCoV-19/Denmark/DCGC-1078/2020\|EPI_ISL_614503\|2020 | ···················· | ···················· | ···················· | ···················· | ···················· | ···················· |
| P | 5132 hCoV-19/Denmark/DCGC-1079/2020\|EPI_ISL_614504\|2020 | ···················· | ···················· | ···················· | ···················· | ···················· | ···················· |
| P | 5133 hCoV-19/Denmark/DCGC-1080/2020\|EPI_ISL_614505\|2020 | ···················· | ···················· | ···················· | ···················· | ···················· | ···················· |
| P | 5134 hCoV-19/Denmark/DCGC-1081/2020\|EPI_ISL_614506\|2020 | ···················· | ···················· | ···················· | ···················· | ···················· | ···················· |
| P | 5135 hCoV-19/Denmark/DCGC-1082/2020\|EPI_ISL_614507\|2020 | ···················· | ···················· | ···················· | ···················· | ···················· | ···················· |
| P | 5136 hCoV-19/Denmark/DCGC-1083/2020\|EPI_ISL_614508\|2020 | ···················· | ···················· | ···················· | ···················· | ···················· | ···················· |
| P | 5137 hCoV-19/Denmark/DCGC-1084/2020\|EPI_ISL_614509\|2020 | ···················· | ···················· | ···················· | ···················· | ···················· | ···················· |
| P | 5138 hCoV-19/Denmark/DCGC-1085/2020\|EPI_ISL_614510\|2020 | ···················· | ···················· | ···················· | ···················· | ···················· | ···················· |
| P | 5139 hCoV-19/Denmark/DCGC-1086/2020\|EPI_ISL_614511\|2020 | ···················· | ···················· | ···················· | ···················· | ···················· | ···················· |
| P | 5140 hCoV-19/Denmark/DCGC-1087/2020\|EPI_ISL_614512\|2020 | ············T······· | ···················· | ···················· | ···················· | ···················· | ···················· |
| P | 5141 hCoV-19/Denmark/DCGC-1088/2020\|EPI_ISL_614513\|2020 | ···················· | ···················· | ···················· | ···················· | ···················· | ···················· |
| P | 5142 hCoV-19/Denmark/DCGC-1089/2020\|EPI_ISL_614514\|2020 | ···················· | ···················· | ···················· | ···················· | ···················· | ···················· |
| P | 5143 hCoV-19/Denmark/DCGC-940/2020\|EPI_ISL_614515\|2020- | ···················· | ···················· | ···················· | ···················· | ···················· | ···················· |
| P | 5144 hCoV-19/Denmark/DCGC-942/2020\|EPI_ISL_614516\|2020- | ···················· | ···················· | ···················· | ···················· | ···················· | ···················· |
| P | 5145 hCoV-19/Denmark/DCGC-4000/2020\|EPI_ISL_615770\|2020 | ···················· | ···················· | ···················· | ···················· | ···················· | ···················· |
| P | 5146 hCoV-19/Denmark/DCGC-4001/2020\|EPI_ISL_615771\|2020 | ···················· | ···················· | ···················· | ···················· | ···················· | ···················· |
| P | 5147 hCoV-19/Denmark/DCGC-4002/2020\|EPI_ISL_615772\|2020 | ···················· | ···················· | ···················· | ···················· | ···················· | ···················· |
| P | 5148 hCoV-19/Denmark/DCGC-6319/2020\|EPI_ISL_615773\|2020 | ···················· | ···················· | ···················· | ···················· | ···················· | ···················· |
| P | 5149 hCoV-19/Denmark/DCGC-4003/2020\|EPI_ISL_615774\|2020 | ···················· | ···················· | ···················· | ···················· | ···················· | ···················· |
| P | 5150 hCoV-19/Denmark/DCGC-4004/2020\|EPI_ISL_615776\|2020 | ···················· | ···················· | ···················· | ···················· | ···················· | ···················· |
| P | 5151 hCoV-19/Denmark/DCGC-4005/2020\|EPI_ISL_615777\|2020 | ···················· | ···················· | ···················· | ···················· | ···················· | ···················· |
| P | 5152 hCoV-19/Denmark/DCGC-4007/2020\|EPI_ISL_615779\|2020 | ···················· | ···················· | ···················· | ···················· | ···················· | ···················· |
| P | 5153 hCoV-19/Denmark/DCGC-4028/2020\|EPI_ISL_615800\|2020 | ···················· | ···················· | ···················· | ···················· | ···················· | ···················· |
| P | 5154 hCoV-19/Denmark/DCGC-4029/2020\|EPI_ISL_615801\|2020 | ···················· | ···················· | ···················· | ···················· | ···················· | ···················· |
| P | 5155 hCoV-19/Denmark/DCGC-4030/2020\|EPI_ISL_615802\|2020 | ···················· | ···················· | ···················· | ···················· | ···················· | ···················· |
| P | 5156 hCoV-19/Denmark/DCGC-6321/2020\|EPI_ISL_615803\|2020 | ···················· | ·A·················· | ···················· | ···················· | ···················· | ···A················ |
| P | 5157 hCoV-19/Denmark/DCGC-4031/2020\|EPI_ISL_615804\|2020 | ···················· | ···················· | ···················· | ···················· | ···················· | ···················· |

| | | | | | | | |
|---|---|---|---|---|---|---|---|
| P | 5158 hCoV-19/Denmark/DCGC-4032/2020\|EPI_ISL_615805\|2020 | ···················· | ···················· | ····················· | ···················· | ···················· | ···················· |
| P | 5159 hCoV-19/Denmark/DCGC-6322/2020\|EPI_ISL_615806\|2020 | ···················· | ···················· | ····················· | ···················· | ···················· | ···················· |
| P | 5160 hCoV-19/Denmark/DCGC-4034/2020\|EPI_ISL_615808\|2020 | ···················· | ···················· | ····················· | ···················· | ···················· | ···················· |
| P | 5161 hCoV-19/Denmark/DCGC-4035/2020\|EPI_ISL_615809\|2020 | ···················· | ···················· | ····················· | ···················· | ···················· | ···················· |
| P | 5162 hCoV-19/Denmark/DCGC-4036/2020\|EPI_ISL_615810\|2020 | ···················· | ···················· | ····················· | ···················· | ···················· | ···················· |
| P | 5163 hCoV-19/Denmark/DCGC-4037/2020\|EPI_ISL_615811\|2020 | ···················· | ···················· | ····················· | ···················· | ···················· | ···················· |
| P | 5164 hCoV-19/Denmark/DCGC-4038/2020\|EPI_ISL_615812\|2020 | ···················· | ···················· | ····················· | ···················· | ···················· | ···················· |
| P | 5165 hCoV-19/Denmark/DCGC-4039/2020\|EPI_ISL_615813\|2020 | ···················· | ···················· | ····················· | ···················· | ···················· | ···················· |
| P | 5166 hCoV-19/Denmark/DCGC-4040/2020\|EPI_ISL_615814\|2020 | ···················· | ···················· | ····················· | ···················· | ···················· | ···················· |
| P | 5167 hCoV-19/Denmark/DCGC-4041/2020\|EPI_ISL_615815\|2020 | ···················· | ···················· | ····················· | ···················· | ···················· | ···················· |
| P | 5168 hCoV-19/Denmark/DCGC-4042/2020\|EPI_ISL_615816\|2020 | ···················· | ···················· | ····················· | ···················· | ···················· | ···················· |
| P | 5169 hCoV-19/Denmark/DCGC-4043/2020\|EPI_ISL_615817\|2020 | ···················· | ···················· | ····················· | ···················· | ···················· | ···················· |
| P | 5170 hCoV-19/Denmark/DCGC-4044/2020\|EPI_ISL_615818\|2020 | ···················· | ···················· | ····················· | ···················· | ···················· | ···················· |
| P | 5171 hCoV-19/Denmark/DCGC-4045/2020\|EPI_ISL_615819\|2020 | ···················· | ···················· | ····················· | ···················· | ···················· | ···················· |
| P | 5172 hCoV-19/Denmark/DCGC-4046/2020\|EPI_ISL_615820\|2020 | ···················· | ···················· | ····················· | ···················· | ···················· | ···················· |
| P | 5173 hCoV-19/Denmark/DCGC-4047/2020\|EPI_ISL_615821\|2020 | ···················· | ···················· | ····················· | ···················· | ···················· | ···················· |
| P | 5174 hCoV-19/Denmark/DCGC-4048/2020\|EPI_ISL_615822\|2020 | ···················· | ···················· | ····················· | ···················· | ···················· | ···················· |
| P | 5175 hCoV-19/Denmark/DCGC-6323/2020\|EPI_ISL_615823\|2020 | ···················· | ···················· | ····················· | ···················· | ···················· | ···················· |
| P | 5176 hCoV-19/Denmark/DCGC-6324/2020\|EPI_ISL_615824\|2020 | ···················· | ···················· | ····················· | ···················· | ···················· | ···················· |
| P | 5177 hCoV-19/Denmark/DCGC-6325/2020\|EPI_ISL_615825\|2020 | ···················· | ···················· | ····················· | ···················· | ···················· | ···················· |
| P | 5178 hCoV-19/Denmark/DCGC-3915/2020\|EPI_ISL_615826\|2020 | ···················· | ···················· | ····················· | ···················· | ···················· | ···················· |
| P | 5179 hCoV-19/Denmark/DCGC-3916/2020\|EPI_ISL_615827\|2020 | ···················· | ···················· | ····················· | ···················· | ···················· | ···················· |
| P | 5180 hCoV-19/Denmark/DCGC-3917/2020\|EPI_ISL_615828\|2020 | ···················· | ···················· | ····················· | ···················· | ···················· | ···················· |
| P | 5181 hCoV-19/Denmark/DCGC-3918/2020\|EPI_ISL_615829\|2020 | ···················· | ···················· | ····················· | ···················· | ···················· | ···················· |
| P | 5182 hCoV-19/Denmark/DCGC-3919/2020\|EPI_ISL_615830\|2020 | ···················· | ···················· | ····················· | ···················· | ···················· | ···················· |
| P | 5183 hCoV-19/Denmark/DCGC-3920/2020\|EPI_ISL_615831\|2020 | ···················· | ···················· | ····················· | ···················· | ···················· | ···················· |
| P | 5184 hCoV-19/Denmark/DCGC-3921/2020\|EPI_ISL_615832\|2020 | ···················· | ···················· | ····················· | ···················· | ···················· | ···················· |
| P | 5185 hCoV-19/Denmark/DCGC-3922/2020\|EPI_ISL_615833\|2020 | ···················· | ···················· | ····················· | ···················· | ···················· | ···················· |
| P | 5186 hCoV-19/Denmark/DCGC-3923/2020\|EPI_ISL_615834\|2020 | ···················· | ···················· | ····················· | ···················· | ···················· | ···················· |
| P | 5187 hCoV-19/Denmark/DCGC-3924/2020\|EPI_ISL_615835\|2020 | ···················· | ···················· | ····················· | ···················· | ···················· | ···················· |
| P | 5188 hCoV-19/Denmark/DCGC-3925/2020\|EPI_ISL_615836\|2020 | ···················· | ···················· | ····················· | ···················· | ···················· | ···················· |
| P | 5189 hCoV-19/Denmark/DCGC-3926/2020\|EPI_ISL_615837\|2020 | ···················· | ···················· | ····················· | ···················· | ···················· | ···················· |
| P | 5190 hCoV-19/Denmark/DCGC-3927/2020\|EPI_ISL_615838\|2020 | ···················· | ···················· | ····················· | ···················· | ···················· | ···················· |
| P | 5191 hCoV-19/Denmark/DCGC-3929/2020\|EPI_ISL_615840\|2020 | ···················· | ·A·················· | ····················· | ···················· | ···················· | ··A················· |
| P | 5192 hCoV-19/Denmark/DCGC-3930/2020\|EPI_ISL_615841\|2020 | ···················· | ···················· | ····················· | ···················· | ···················· | ···················· |
| P | 5193 hCoV-19/Denmark/DCGC-3931/2020\|EPI_ISL_615842\|2020 | ···················· | ···················· | ····················· | ···················· | ···················· | ···················· |
| P | 5194 hCoV-19/Denmark/DCGC-3933/2020\|EPI_ISL_615844\|2020 | ···················· | ···················· | ····················· | ···················· | ···················· | ···················· |
| P | 5195 hCoV-19/Denmark/DCGC-3934/2020\|EPI_ISL_615845\|2020 | ···················· | ···················· | ····················· | ···················· | ···················· | ···················· |
| P | 5196 hCoV-19/Denmark/DCGC-3935/2020\|EPI_ISL_615846\|2020 | ···················· | ···················· | ····················· | ···················· | ···················· | ···················· |
| P | 5197 hCoV-19/Denmark/DCGC-3936/2020\|EPI_ISL_615847\|2020 | ···················· | ···················· | ····················· | ···················· | ···················· | ···················· |
| P | 5198 hCoV-19/Denmark/DCGC-1263/2020\|EPI_ISL_618040\|2020 | ···················· | ···················· | ····················· | ···················· | ···················· | ···················· |
| P | 5199 hCoV-19/Denmark/DCGC-1264/2020\|EPI_ISL_618041\|2020 | ···················· | ···················· | ····················· | ···················· | ···················· | ···················· |
| P | 5200 hCoV-19/Denmark/DCGC-1265/2020\|EPI_ISL_618043\|2020 | ···················· | ···················· | ····················· | ···················· | ···················· | ···················· |
| P | 5201 hCoV-19/Denmark/DCGC-1266/2020\|EPI_ISL_618045\|2020 | ···················· | ···················· | ····················· | ···················· | ···················· | ···················· |
| P | 5203 hCoV-19/Denmark/DCGC-1313/2020\|EPI_ISL_618047\|2020 | ···················· | ···················· | ····················· | ···················· | ···················· | ···················· |
| P | 5204 hCoV-19/Denmark/DCGC-1314/2020\|EPI_ISL_618048\|2020 | ···················· | ···················· | ····················· | ···················· | ···················· | ···················· |
| P | 5205 hCoV-19/Denmark/DCGC-1315/2020\|EPI_ISL_618049\|2020 | ···················· | ···················· | ····················· | ···················· | ···················· | ···················· |
| P | 5206 hCoV-19/Denmark/DCGC-1316/2020\|EPI_ISL_618050\|2020 | ···················· | ···················· | ····················· | ···················· | ···················· | ···················· |
| P | 5207 hCoV-19/Denmark/DCGC-1317/2020\|EPI_ISL_618051\|2020 | ···················· | ···················· | ····················· | ···················· | ···················· | ···················· |
| P | 5208 hCoV-19/Denmark/DCGC-1318/2020\|EPI_ISL_618052\|2020 | ···················· | ···················· | ····················· | ···················· | ···················· | ···················· |
| P | 5209 hCoV-19/Denmark/DCGC-1319/2020\|EPI_ISL_618053\|2020 | ···················· | ···················· | ····················· | ···················· | ···················· | ···················· |
| P | 5210 hCoV-19/Denmark/DCGC-1320/2020\|EPI_ISL_618054\|2020 | ···················· | ···················· | ····················· | ···················· | ···················· | ···················· |
| P | 5211 hCoV-19/Denmark/DCGC-2069/2020\|EPI_ISL_618055\|2020 | ···················· | ···················· | ····················· | ···················· | ···················· | ···················· |
| P | 5212 hCoV-19/Denmark/DCGC-1267/2020\|EPI_ISL_618056\|2020 | ···················· | ···················· | ····················· | ···················· | ···················· | ···················· |
| P | 5213 hCoV-19/Denmark/DCGC-1268/2020\|EPI_ISL_618057\|2020 | ···················· | ···················· | ····················· | ···················· | ···················· | ···················· |
| P | 5214 hCoV-19/Denmark/DCGC-1269/2020\|EPI_ISL_618058\|2020 | ···················· | ···················· | ····················· | ···················· | ···················· | ···················· |
| P | 5216 hCoV-19/Denmark/DCGC-1270/2020\|EPI_ISL_618062\|2020 | ···················· | ···················· | ····················· | ···················· | ···················· | ···················· |
| P | 5218 hCoV-19/Denmark/DCGC-1271/2020\|EPI_ISL_618064\|2020 | ···················· | ···················· | ····················· | ···················· | ···················· | ···················· |
| P | 5219 hCoV-19/Denmark/DCGC-1272/2020\|EPI_ISL_618065\|2020 | ···················· | ···················· | ····················· | ···················· | ···················· | ···················· |
| P | 5220 hCoV-19/Denmark/DCGC-1273/2020\|EPI_ISL_618066\|2020 | ···················· | ···················· | ····················· | ···················· | ···················· | ···················· |
| P | 5221 hCoV-19/Denmark/DCGC-1274/2020\|EPI_ISL_618067\|2020 | ···················· | ···················· | ····················· | ···················· | ···················· | ···················· |
| P | 5222 hCoV-19/Denmark/DCGC-1275/2020\|EPI_ISL_618068\|2020 | ···················· | ···················· | ····················· | ···················· | ···················· | ···················· |
| P | 5223 hCoV-19/Denmark/DCGC-1276/2020\|EPI_ISL_618069\|2020 | ···················· | ···················· | ····················· | ···················· | ···················· | ···················· |
| P | 5224 hCoV-19/Denmark/DCGC-1277/2020\|EPI_ISL_618070\|2020 | ···················· | ···················· | ····················· | ···················· | ···················· | ···················· |
| P | 5225 hCoV-19/Denmark/DCGC-1278/2020\|EPI_ISL_618071\|2020 | ···················· | ···················· | ····················· | ···················· | ···················· | ···················· |
| P | 5226 hCoV-19/Denmark/DCGC-1820/2020\|EPI_ISL_618072\|2020 | ···················· | ···················· | ····················· | ···················· | ···················· | ···················· |
| P | 5227 hCoV-19/Denmark/DCGC-1821/2020\|EPI_ISL_618073\|2020 | ···················· | ···················· | ····················· | ···················· | ···················· | ···················· |
| P | 5228 hCoV-19/Denmark/DCGC-1351/2020\|EPI_ISL_618074\|2020 | ···················· | ···················· | ····················· | ···················· | ···················· | ···················· |
| P | 5229 hCoV-19/Denmark/DCGC-1292/2020\|EPI_ISL_618076\|2020 | ···················· | ···················· | ····················· | ···················· | ···················· | ···················· |
| P | 5230 hCoV-19/Denmark/DCGC-1293/2020\|EPI_ISL_618077\|2020 | ···················· | ···················· | ····················· | ···················· | ···················· | ···················· |
| P | 5231 hCoV-19/Denmark/DCGC-1294/2020\|EPI_ISL_618078\|2020 | ···················· | ···················· | ····················· | ···················· | ···················· | ···················· |
| P | 5232 hCoV-19/Denmark/DCGC-1296/2020\|EPI_ISL_618079\|2020 | ···················· | ···················· | ····················· | ···················· | ···················· | ···················· |
| P | 5233 hCoV-19/Denmark/DCGC-1297/2020\|EPI_ISL_618080\|2020 | ···················· | ···················· | ····················· | ···················· | ···················· | ···················· |
| P | 5234 hCoV-19/Denmark/DCGC-1298/2020\|EPI_ISL_618081\|2020 | ···················· | ···················· | ····················· | ···················· | ···················· | ···················· |
| P | 5235 hCoV-19/Denmark/DCGC-1299/2020\|EPI_ISL_618082\|2020 | ···················· | ···················· | ····················· | ···················· | ···················· | ···················· |
| P | 5236 hCoV-19/Denmark/DCGC-1300/2020\|EPI_ISL_618083\|2020 | ···················· | ···················· | ····················· | ···················· | ···················· | ···················· |
| P | 5237 hCoV-19/Denmark/DCGC-1301/2020\|EPI_ISL_618084\|2020 | ···················· | ···················· | ····················· | ···················· | ···················· | ···················· |
| P | 5238 hCoV-19/Denmark/DCGC-1302/2020\|EPI_ISL_618085\|2020 | ···················· | ···················· | ····················· | ···················· | ···················· | ···················· |
| P | 5239 hCoV-19/Denmark/DCGC-1303/2020\|EPI_ISL_618086\|2020 | ···················· | ···················· | ····················· | ···················· | ···················· | ···················· |
| P | 5240 hCoV-19/Denmark/DCGC-1304/2020\|EPI_ISL_618087\|2020 | ···················· | ···················· | ····················· | ···················· | ···················· | ···················· |

| | | | | | | | | |
|---|---|---|---|---|---|---|---|---|
| P | 5241 | hCoV-19/Denmark/DCGC-1305/2020\|EPI_ISL_618088\|2020 | ···················· | ···················· | ···················· | ···················· | ···················· | ···················· |
| P | 5242 | hCoV-19/Denmark/DCGC-1306/2020\|EPI_ISL_618089\|2020 | ···················· | ···················· | ···················· | ···················· | ···················· | ···················· |
| P | 5243 | hCoV-19/Denmark/DCGC-1307/2020\|EPI_ISL_618090\|2020 | ···················· | ···················· | ···················· | ···················· | ···················· | ···················· |
| P | 5244 | hCoV-19/Denmark/DCGC-1308/2020\|EPI_ISL_618091\|2020 | ···················· | ···················· | ···················· | ···················· | ···················· | ···················· |
| P | 5245 | hCoV-19/Denmark/DCGC-1451/2020\|EPI_ISL_618092\|2020 | ···················· | ···················· | ···················· | ···················· | ···················· | ···················· |
| P | 5246 | hCoV-19/Denmark/DCGC-1309/2020\|EPI_ISL_618093\|2020 | ···················· | ···················· | ···················· | ···················· | ···················· | ···················· |
| P | 5247 | hCoV-19/Denmark/DCGC-1452/2020\|EPI_ISL_618094\|2020 | ···················· | ···················· | ···················· | ···················· | ···················· | ···················· |
| P | 5248 | hCoV-19/Denmark/DCGC-1310/2020\|EPI_ISL_618095\|2020 | ···················· | ···················· | ···················· | ···················· | ···················· | ···················· |
| P | 5249 | hCoV-19/Denmark/DCGC-1312/2020\|EPI_ISL_618096\|2020 | ···················· | ···················· | ···················· | ···················· | ···················· | ···················· |
| P | 5250 | hCoV-19/Denmark/DCGC-1280/2020\|EPI_ISL_618097\|2020 | ···················· | ···················· | ···················· | ···················· | ···················· | ···················· |
| P | 5251 | hCoV-19/Denmark/DCGC-1450/2020\|EPI_ISL_618098\|2020 | ···················· | ···················· | ···················· | ···················· | ···················· | ···················· |
| P | 5252 | hCoV-19/Denmark/DCGC-1279/2020\|EPI_ISL_618099\|2020 | ···················· | ···················· | ···················· | ···················· | ···················· | ···················· |
| P | 5253 | hCoV-19/Denmark/DCGC-7324/2020\|EPI_ISL_619310\|2020 | ···················· | ···················· | ···················· | ···················· | ···················· | ···················· |
| P | 5254 | hCoV-19/Denmark/DCGC-7330/2020\|EPI_ISL_619311\|2020 | ···················· | ···················· | ···················· | ···················· | ···················· | ···················· |
| P | 5255 | hCoV-19/Denmark/DCGC-7346/2020\|EPI_ISL_619312\|2020 | ···················· | ·A·················· | ···················· | ···················· | ···················· | ··A················· |
| P | 5256 | hCoV-19/Denmark/DCGC-7373/2020\|EPI_ISL_619313\|2020 | ···················· | ···················· | ···················· | ···················· | ···················· | ···················· |
| P | 5257 | hCoV-19/Denmark/DCGC-7319/2020\|EPI_ISL_619314\|2020 | ···················· | ···················· | ···················· | ···················· | ···················· | ···················· |
| P | 5258 | hCoV-19/Denmark/DCGC-7325/2020\|EPI_ISL_619315\|2020 | ···················· | ···················· | ···················· | ···················· | ···················· | ···················· |
| P | 5259 | hCoV-19/Denmark/DCGC-7331/2020\|EPI_ISL_619316\|2020 | ···················· | ·A·················· | ···················· | ···················· | ···················· | ···················· |
| P | 5260 | hCoV-19/Denmark/DCGC-7339/2020\|EPI_ISL_619317\|2020 | ···················· | ···················· | ···················· | ···················· | ···················· | ···················· |
| P | 5261 | hCoV-19/Denmark/DCGC-1613/2020\|EPI_ISL_619318\|2020 | ···················· | ···················· | ···················· | ···················· | ···················· | ···················· |
| P | 5262 | hCoV-19/Denmark/DCGC-7353/2020\|EPI_ISL_619319\|2020 | ···················· | ···················· | ···················· | ···················· | ···················· | ···················· |
| P | 5263 | hCoV-19/Denmark/DCGC-7359/2020\|EPI_ISL_619320\|2020 | ···················· | ···················· | ···················· | ···················· | ···················· | ···················· |
| P | 5264 | hCoV-19/Denmark/DCGC-2045/2020\|EPI_ISL_619321\|2020 | ···················· | ···················· | ···················· | ···················· | ···················· | ···················· |
| P | 5265 | hCoV-19/Denmark/DCGC-7411/2020\|EPI_ISL_619322\|2020 | ···················· | ···················· | ···················· | ···················· | ···················· | ···················· |
| P | 5266 | hCoV-19/Denmark/DCGC-7426/2020\|EPI_ISL_619323\|2020 | ···················· | ···················· | ···················· | ···················· | ···················· | ···················· |
| P | 5267 | hCoV-19/Denmark/DCGC-7434/2020\|EPI_ISL_619324\|2020 | ···················· | ···················· | ···················· | ···················· | ···················· | ···················· |
| P | 5268 | hCoV-19/Denmark/DCGC-7438/2020\|EPI_ISL_619325\|2020 | ···················· | ···················· | ···················· | ···················· | ···················· | ···················· |
| P | 5269 | hCoV-19/Denmark/DCGC-7446/2020\|EPI_ISL_619326\|2020 | ···················· | ···················· | ···················· | ···················· | ···················· | ···················· |
| P | 5270 | hCoV-19/Denmark/DCGC-7392/2020\|EPI_ISL_619327\|2020 | ···················· | ···················· | ···················· | ···················· | ···················· | ···················· |
| P | 5271 | hCoV-19/Denmark/DCGC-1563/2020\|EPI_ISL_619328\|2020 | ···················· | ···················· | ···················· | ···················· | ···················· | ···················· |
| P | 5272 | hCoV-19/Denmark/DCGC-7420/2020\|EPI_ISL_619329\|2020 | ···················· | ·A·················· | ···················· | ···················· | ···················· | ···················· |
| P | 5273 | hCoV-19/Denmark/DCGC-7461/2020\|EPI_ISL_619370\|2020 | ···················· | ···················· | ···················· | ···················· | ···················· | ···················· |
| P | 5274 | hCoV-19/Denmark/DCGC-7475/2020\|EPI_ISL_619371\|2020 | ···················· | ···················· | ···················· | ···················· | ···················· | ···················· |
| P | 5275 | hCoV-19/Denmark/DCGC-7485/2020\|EPI_ISL_619372\|2020 | ···················· | ···················· | ···················· | ···················· | ···················· | ···················· |
| P | 5276 | hCoV-19/Denmark/DCGC-7500/2020\|EPI_ISL_619373\|2020 | ···················· | ···················· | ···················· | ···················· | ···················· | ···················· |
| P | 5277 | hCoV-19/Denmark/DCGC-7509/2020\|EPI_ISL_619374\|2020 | ···················· | ···················· | ···················· | ···················· | ···················· | ···················· |
| P | 5278 | hCoV-19/Denmark/DCGC-7522/2020\|EPI_ISL_619375\|2020 | ···················· | ···················· | ···················· | ···················· | ···················· | ···················· |
| P | 5279 | hCoV-19/Denmark/DCGC-7469/2020\|EPI_ISL_619376\|2020 | ···················· | ···················· | ···················· | ···················· | ···················· | ···················· |
| P | 5280 | hCoV-19/Denmark/DCGC-7476/2020\|EPI_ISL_619377\|2020 | ···················· | ···················· | ···················· | ···················· | ···················· | ···················· |
| P | 5281 | hCoV-19/Denmark/DCGC-1610/2020\|EPI_ISL_619378\|2020 | ···················· | ···················· | ···················· | ···················· | ···················· | ···················· |
| P | 5282 | hCoV-19/Denmark/DCGC-7479/2020\|EPI_ISL_619379\|2020 | ···················· | ···················· | ···················· | ···················· | ···················· | ···················· |
| P | 5283 | hCoV-19/Denmark/DCGC-7486/2020\|EPI_ISL_619380\|2020 | ···················· | ···················· | ···················· | ···················· | ···················· | ···················· |
| P | 5284 | hCoV-19/Denmark/DCGC-7501/2020\|EPI_ISL_619381\|2020 | ···················· | ···················· | ···················· | ···················· | ···················· | ···················· |
| P | 5285 | hCoV-19/Denmark/DCGC-7516/2020\|EPI_ISL_619382\|2020 | ···················· | ···················· | ···················· | ···················· | ···················· | ···················· |
| P | 5286 | hCoV-19/Denmark/DCGC-7463/2020\|EPI_ISL_619383\|2020 | ···················· | ···················· | ···················· | ···················· | ···················· | ···················· |
| P | 5287 | hCoV-19/Denmark/DCGC-1609/2020\|EPI_ISL_619384\|2020 | ···················· | ···················· | ···················· | ···················· | ···················· | ···················· |
| P | 5288 | hCoV-19/Denmark/DCGC-7487/2020\|EPI_ISL_619385\|2020 | ···················· | ···················· | ···················· | ···················· | ···················· | ···················· |
| P | 5289 | hCoV-19/Denmark/DCGC-7494/2020\|EPI_ISL_619386\|2020 | ···················· | ···················· | ···················· | ···················· | ···················· | ···················· |
| P | 5290 | hCoV-19/Denmark/DCGC-7523/2020\|EPI_ISL_619387\|2020 | ···················· | ···················· | ···················· | ···················· | ···················· | ···················· |
| P | 5291 | hCoV-19/Denmark/DCGC-7457/2020\|EPI_ISL_619388\|2020 | ···················· | ···················· | ···················· | ···················· | ···················· | ···················· |
| P | 5292 | hCoV-19/Denmark/DCGC-2047/2020\|EPI_ISL_619389\|2020 | ···················· | ···················· | ···················· | ···················· | ···················· | ···················· |
| P | 5293 | hCoV-19/Denmark/DCGC-7477/2020\|EPI_ISL_619390\|2020 | ···················· | ···················· | ···················· | ···················· | ···················· | ···················· |
| P | 5294 | hCoV-19/Denmark/DCGC-7488/2020\|EPI_ISL_619391\|2020 | ···················· | ···················· | ···················· | ···················· | ···················· | ···················· |
| P | 5295 | hCoV-19/Denmark/DCGC-7495/2020\|EPI_ISL_619392\|2020 | ···················· | ···················· | ···················· | ···················· | ···················· | ···················· |
| P | 5296 | hCoV-19/Denmark/DCGC-7458/2020\|EPI_ISL_619393\|2020 | ···················· | ···················· | ···················· | ···················· | ···················· | ···················· |
| P | 5297 | hCoV-19/Denmark/DCGC-7471/2020\|EPI_ISL_619394\|2020 | ···················· | ···················· | ···················· | ···················· | ···················· | ···················· |
| P | 5298 | hCoV-19/Denmark/DCGC-7481/2020\|EPI_ISL_619395\|2020 | ···················· | ···················· | ···················· | ···················· | ···················· | ···················· |
| P | 5299 | hCoV-19/Denmark/DCGC-7489/2020\|EPI_ISL_619396\|2020 | ···················· | ···················· | ···················· | ···················· | ···················· | ···················· |
| P | 5300 | hCoV-19/Denmark/DCGC-7496/2020\|EPI_ISL_619397\|2020 | ···················· | ···················· | ···················· | ···················· | ···················· | ···················· |
| P | 5301 | hCoV-19/Denmark/DCGC-1608/2020\|EPI_ISL_619398\|2020 | ···················· | ···················· | ···················· | ···················· | ···················· | ···················· |
| P | 5302 | hCoV-19/Denmark/DCGC-7519/2020\|EPI_ISL_619399\|2020 | ···················· | ···················· | ···················· | ···················· | ···················· | ···················· |
| P | 5303 | hCoV-19/Denmark/DCGC-6105/2020\|EPI_ISL_620300\|2020 | ···················· | ···················· | ···················· | ···················· | ···················· | ···················· |
| P | 5304 | hCoV-19/Denmark/DCGC-6106/2020\|EPI_ISL_620301\|2020 | ···················· | ···················· | ···················· | ···················· | ···················· | ···················· |
| P | 5305 | hCoV-19/Denmark/DCGC-6403/2020\|EPI_ISL_620303\|2020 | ···················· | ···················· | ···················· | ···················· | ···················· | ···················· |
| P | 5306 | hCoV-19/Denmark/DCGC-6405/2020\|EPI_ISL_620305\|2020 | ···················· | ···················· | ···················· | ···················· | ···················· | ···················· |
| P | 5307 | hCoV-19/Denmark/DCGC-6406/2020\|EPI_ISL_620306\|2020 | ···················· | ···················· | ···················· | ···················· | ···················· | ···················· |
| P | 5308 | hCoV-19/Denmark/DCGC-6407/2020\|EPI_ISL_620307\|2020 | ···················· | ·A·················· | ···················· | ···················· | ···················· | ···················· |
| P | 5309 | hCoV-19/Denmark/DCGC-6410/2020\|EPI_ISL_620310\|2020 | ···················· | ···················· | ···················· | ···················· | ···················· | ···················· |
| P | 5310 | hCoV-19/Denmark/DCGC-6411/2020\|EPI_ISL_620311\|2020 | ···················· | ···················· | ···················· | ···················· | ···················· | ···················· |
| P | 5311 | hCoV-19/Denmark/DCGC-6412/2020\|EPI_ISL_620312\|2020 | ···················· | ···················· | ···················· | ···················· | ···················· | ···················· |
| P | 5312 | hCoV-19/Denmark/DCGC-6413/2020\|EPI_ISL_620313\|2020 | ···················· | ···················· | ···················· | ···················· | ···················· | ···················· |
| P | 5313 | hCoV-19/Denmark/DCGC-6458/2020\|EPI_ISL_620358\|2020 | ···················· | ···················· | ···················· | ···················· | ···················· | ···················· |
| P | 5314 | hCoV-19/Denmark/DCGC-6459/2020\|EPI_ISL_620359\|2020 | ···················· | ···················· | ···················· | ···················· | ···················· | ···················· |
| P | 5315 | hCoV-19/Denmark/DCGC-6460/2020\|EPI_ISL_620360\|2020 | ···················· | ···················· | ···················· | ···················· | ···················· | ···················· |
| P | 5316 | hCoV-19/Denmark/DCGC-6461/2020\|EPI_ISL_620361\|2020 | ···················· | ···················· | ···················· | ···················· | ···················· | ···················· |
| P | 5317 | hCoV-19/Denmark/DCGC-6462/2020\|EPI_ISL_620362\|2020 | ···················· | ···················· | ···················· | ···················· | ···················· | ···················· |
| P | 5318 | hCoV-19/Denmark/DCGC-6463/2020\|EPI_ISL_620363\|2020 | ···················· | ···················· | ···················· | ···················· | ···················· | ···················· |
| P | 5319 | hCoV-19/Denmark/DCGC-6464/2020\|EPI_ISL_620364\|2020 | ···················· | ···················· | ···················· | ···················· | ···················· | ···················· |
| P | 5320 | hCoV-19/Denmark/DCGC-6465/2020\|EPI_ISL_620365\|2020 | ···················· | ···················· | ···················· | ···················· | ···················· | ···················· |

| | | | | | | | |
|---|---|---|---|---|---|---|---|
| P | 5321 hCoV-19/Denmark/DCGC-6466/2020\|EPI_ISL_620366\|2020 | ···················· | ···················· | ····················· | ···················· | ···················· | ···················· |
| P | 5322 hCoV-19/Denmark/DCGC-6467/2020\|EPI_ISL_620367\|2020 | ···················· | ···················· | ····················· | ···················· | ···················· | ···················· |
| P | 5323 hCoV-19/Denmark/DCGC-6468/2020\|EPI_ISL_620368\|2020 | ···················· | ···················· | ····················· | ···················· | ···················· | ···················· |
| P | 5324 hCoV-19/Denmark/DCGC-6469/2020\|EPI_ISL_620369\|2020 | ···················· | ···················· | ····················· | ···················· | ···················· | ···················· |
| P | 5325 hCoV-19/Denmark/DCGC-6470/2020\|EPI_ISL_620370\|2020 | ···················· | ·A·················· | ····················· | ···················· | ···················· | ···················· |
| P | 5326 hCoV-19/Denmark/DCGC-6471/2020\|EPI_ISL_620371\|2020 | ···················· | ···················· | ····················· | ···················· | ···················· | ···················· |
| P | 5327 hCoV-19/Denmark/DCGC-6472/2020\|EPI_ISL_620372\|2020 | ···················· | ·A·················· | ····················· | ···················· | ···················· | ··A················· |
| P | 5328 hCoV-19/Denmark/DCGC-6474/2020\|EPI_ISL_620374\|2020 | ···················· | ···················· | ····················· | ···················· | ···················· | ···················· |
| P | 5329 hCoV-19/Denmark/DCGC-6476/2020\|EPI_ISL_620376\|2020 | ···················· | ···················· | ····················· | ···················· | ···················· | ···················· |
| P | 5330 hCoV-19/Denmark/DCGC-6477/2020\|EPI_ISL_620377\|2020 | ···················· | ···················· | ····················· | ···················· | ···················· | ···················· |
| P | 5331 hCoV-19/Denmark/DCGC-6478/2020\|EPI_ISL_620378\|2020 | ···················· | ·A·················· | ····················· | ···················· | ···················· | ··A················· |
| P | 5332 hCoV-19/Denmark/DCGC-6479/2020\|EPI_ISL_620379\|2020 | ···················· | ···················· | ····················· | ···················· | ···················· | ···················· |
| P | 5333 hCoV-19/Denmark/DCGC-6480/2020\|EPI_ISL_620380\|2020 | ···················· | ···················· | ····················· | ···················· | ···················· | ···················· |
| P | 5334 hCoV-19/Denmark/DCGC-6481/2020\|EPI_ISL_620381\|2020 | ···················· | ·A·················· | ····················· | ···················· | ···················· | ···················· |
| P | 5335 hCoV-19/Denmark/DCGC-6483/2020\|EPI_ISL_620383\|2020 | ···················· | ···················· | ····················· | ···················· | ···················· | ···················· |
| P | 5336 hCoV-19/Denmark/DCGC-6484/2020\|EPI_ISL_620384\|2020 | ···················· | ···················· | ····················· | ···················· | ···················· | ···················· |
| P | 5337 hCoV-19/Denmark/DCGC-6485/2020\|EPI_ISL_620385\|2020 | ···················· | ···················· | ····················· | ···················· | ···················· | ···················· |
| P | 5338 hCoV-19/Denmark/DCGC-6486/2020\|EPI_ISL_620386\|2020 | ···················· | ·A·················· | ····················· | ···················· | ···················· | ··A················· |
| P | 5339 hCoV-19/Denmark/DCGC-6489/2020\|EPI_ISL_620389\|2020 | ···················· | ···················· | ····················· | ···················· | ···················· | ···················· |
| P | 5340 hCoV-19/Denmark/DCGC-6491/2020\|EPI_ISL_620391\|2020 | ···················· | ···················· | ····················· | ···················· | ···················· | ···················· |
| P | 5341 hCoV-19/Denmark/DCGC-6492/2020\|EPI_ISL_620392\|2020 | ···················· | ···················· | ····················· | ···················· | ···················· | ···················· |
| P | 5342 hCoV-19/Denmark/DCGC-6493/2020\|EPI_ISL_620393\|2020 | ···················· | ···················· | ····················· | ···················· | ···················· | ···················· |
| P | 5343 hCoV-19/Denmark/DCGC-6496/2020\|EPI_ISL_620396\|2020 | ···················· | ·A·················· | ····················· | ···················· | ···················· | ···················· |
| P | 5344 hCoV-19/Denmark/DCGC-6497/2020\|EPI_ISL_620397\|2020 | ···················· | ···················· | ····················· | ···················· | ···················· | ···················· |
| P | 5345 hCoV-19/Denmark/DCGC-6498/2020\|EPI_ISL_620398\|2020 | ···················· | ···················· | ····················· | ···················· | ···················· | ···················· |
| P | 5346 hCoV-19/Denmark/DCGC-6499/2020\|EPI_ISL_620399\|2020 | ···················· | ···················· | ····················· | ···················· | ···················· | ···················· |
| P | 5347 hCoV-19/Denmark/DCGC-5085/2020\|EPI_ISL_621623\|2020 | ···················· | ···················· | ····················· | ···················· | ···················· | ···················· |
| P | 5348 hCoV-19/Denmark/DCGC-5086/2020\|EPI_ISL_621624\|2020 | ···················· | ···················· | ····················· | ···················· | ···················· | ···················· |
| P | 5349 hCoV-19/Denmark/DCGC-5087/2020\|EPI_ISL_621625\|2020 | ···················· | ···················· | ····················· | ···················· | ···················· | ···················· |
| P | 5350 hCoV-19/Denmark/DCGC-5088/2020\|EPI_ISL_621626\|2020 | ···················· | ···················· | ····················· | ···················· | ···················· | ···················· |
| P | 5351 hCoV-19/Denmark/DCGC-5089/2020\|EPI_ISL_621627\|2020 | ···················· | ···················· | ····················· | ···················· | ···················· | ···················· |
| P | 5352 hCoV-19/Denmark/DCGC-5090/2020\|EPI_ISL_621628\|2020 | ···················· | ···················· | ····················· | ···················· | ···················· | ···················· |
| P | 5353 hCoV-19/Denmark/DCGC-5091/2020\|EPI_ISL_621629\|2020 | ···················· | ···················· | ····················· | ···················· | ···················· | ···················· |
| P | 5354 hCoV-19/Denmark/DCGC-5093/2020\|EPI_ISL_621631\|2020 | ···················· | ···················· | ····················· | ···················· | ···················· | ···················· |
| P | 5355 hCoV-19/Denmark/DCGC-5094/2020\|EPI_ISL_621632\|2020 | ···················· | ···················· | ····················· | ···················· | ···················· | ···················· |
| P | 5356 hCoV-19/Denmark/DCGC-5095/2020\|EPI_ISL_621633\|2020 | ···················· | ···················· | ····················· | ···················· | ···················· | ···················· |
| P | 5357 hCoV-19/Denmark/DCGC-5096/2020\|EPI_ISL_621634\|2020 | ···················· | ···················· | ····················· | ···················· | ···················· | ···················· |
| P | 5358 hCoV-19/Denmark/DCGC-5097/2020\|EPI_ISL_621635\|2020 | ···················· | ···················· | ····················· | ···················· | ···················· | ···················· |
| P | 5359 hCoV-19/Denmark/DCGC-5098/2020\|EPI_ISL_621636\|2020 | ···················· | ···················· | ····················· | ···················· | ···················· | ···················· |
| P | 5360 hCoV-19/Denmark/DCGC-5100/2020\|EPI_ISL_621638\|2020 | ···················· | ···················· | ····················· | ···················· | ···················· | ···················· |
| P | 5361 hCoV-19/Denmark/DCGC-5102/2020\|EPI_ISL_621640\|2020 | ···················· | ···················· | ····················· | ···················· | ···················· | ···················· |
| P | 5362 hCoV-19/Denmark/DCGC-5103/2020\|EPI_ISL_621641\|2020 | ···················· | ···················· | ····················· | ···················· | ···················· | ···················· |
| P | 5363 hCoV-19/Denmark/DCGC-5104/2020\|EPI_ISL_621642\|2020 | ···················· | ···················· | ····················· | ···················· | ···················· | ···················· |
| P | 5364 hCoV-19/Denmark/DCGC-5106/2020\|EPI_ISL_621644\|2020 | ···················· | ···················· | ····················· | ···················· | ···················· | ···················· |
| P | 5365 hCoV-19/Denmark/DCGC-5151/2020\|EPI_ISL_621689\|2020 | ···················· | ···················· | ····················· | ···················· | ···················· | ···················· |
| P | 5366 hCoV-19/Denmark/DCGC-5152/2020\|EPI_ISL_621690\|2020 | ···················· | ···················· | ····················· | ···················· | ···················· | ···················· |
| P | 5367 hCoV-19/Denmark/DCGC-5153/2020\|EPI_ISL_621691\|2020 | ···················· | ···················· | ····················· | ···················· | ···················· | ···················· |
| P | 5368 hCoV-19/Denmark/DCGC-5154/2020\|EPI_ISL_621692\|2020 | ···················· | ···················· | ····················· | ···················· | ···················· | ···················· |
| P | 5369 hCoV-19/Denmark/DCGC-5155/2020\|EPI_ISL_621693\|2020 | ···················· | ···················· | ····················· | ···················· | ···················· | ···················· |
| P | 5370 hCoV-19/Denmark/DCGC-5156/2020\|EPI_ISL_621694\|2020 | ···················· | ···················· | ····················· | ···················· | ···················· | ···················· |
| P | 5371 hCoV-19/Denmark/DCGC-5157/2020\|EPI_ISL_621695\|2020 | ···················· | ···················· | ····················· | ···················· | ···················· | ···················· |
| P | 5372 hCoV-19/Denmark/DCGC-5159/2020\|EPI_ISL_621697\|2020 | ···················· | ···················· | ····················· | ···················· | ···················· | ···················· |
| P | 5373 hCoV-19/Denmark/DCGC-5160/2020\|EPI_ISL_621698\|2020 | ···················· | ···················· | ····················· | ···················· | ···················· | ···················· |
| P | 5374 hCoV-19/Denmark/DCGC-5161/2020\|EPI_ISL_621699\|2020 | ···················· | ···················· | ····················· | ···················· | ···················· | ···················· |
| P | 5375 hCoV-19/England/ALDP-AA5425/2020\|EPI_ISL_625102\|20 | ···················· | ···················· | ····················· | ···················· | ···················· | ···················· |
| P | 5376 hCoV-19/England/ALDP-AA0A73/2020\|EPI_ISL_625103\|20 | ···················· | ···················· | ····················· | ···················· | ···················· | ···················· |
| P | 5377 hCoV-19/Denmark/DCGC-845/2020\|EPI_ISL_614419\|2020- | ···················· | ···················· | ····················· | ···················· | ···················· | ···················· |
| P | 5378 hCoV-19/Denmark/DCGC-901/2020\|EPI_ISL_614427\|2020- | ···················· | ···················· | ····················· | ···················· | ···················· | ···················· |
| P | 5379 hCoV-19/Denmark/DCGC-1108/2020\|EPI_ISL_614428\|2020 | ···················· | ···················· | ····················· | ···················· | ···················· | ···················· |
| P | 5380 hCoV-19/Denmark/DCGC-1110/2020\|EPI_ISL_614429\|2020 | ···················· | ···················· | ····················· | ···················· | ···················· | ···················· |
| P | 5381 hCoV-19/Denmark/DCGC-1127/2020\|EPI_ISL_614430\|2020 | ···················· | ···················· | ····················· | ···················· | ···················· | ···················· |
| P | 5382 hCoV-19/Denmark/DCGC-1106/2020\|EPI_ISL_614431\|2020 | ···················· | ···················· | ····················· | ···················· | ···················· | ···················· |
| P | 5383 hCoV-19/Denmark/DCGC-862/2020\|EPI_ISL_614432\|2020- | ···················· | ···················· | ····················· | ···················· | ···················· | ···················· |
| P | 5384 hCoV-19/Denmark/DCGC-843/2020\|EPI_ISL_614433\|2020- | ···················· | ···················· | ····················· | ···················· | ···················· | ···················· |
| P | 5385 hCoV-19/Denmark/DCGC-1004/2020\|EPI_ISL_614434\|2020 | ···················· | ···················· | ····················· | ···················· | ···················· | ···················· |
| P | 5386 hCoV-19/Denmark/DCGC-947/2020\|EPI_ISL_614435\|2020- | ···················· | ···················· | ····················· | ···················· | ···················· | ···················· |
| P | 5387 hCoV-19/Denmark/DCGC-984/2020\|EPI_ISL_614436\|2020- | ···················· | ···················· | ····················· | ···················· | ···················· | ···················· |
| P | 5388 hCoV-19/Denmark/DCGC-978/2020\|EPI_ISL_614437\|2020- | ···················· | ···················· | ····················· | ···················· | ···················· | ···················· |
| P | 5389 hCoV-19/Denmark/DCGC-968/2020\|EPI_ISL_614438\|2020- | ···················· | ···················· | ····················· | ···················· | ···················· | ···················· |
| P | 5390 hCoV-19/Denmark/DCGC-1097/2020\|EPI_ISL_614439\|2020 | ···················· | ···················· | ····················· | ···················· | ···················· | ···················· |
| P | 5391 hCoV-19/Denmark/DCGC-779/2020\|EPI_ISL_614450\|2020- | ···················· | ···················· | ····················· | ···················· | ···················· | ···················· |
| P | 5392 hCoV-19/Denmark/DCGC-887/2020\|EPI_ISL_614451\|2020- | ···················· | ···················· | ····················· | ···················· | ···················· | ···················· |
| P | 5393 hCoV-19/Denmark/DCGC-888/2020\|EPI_ISL_614452\|2020- | ···················· | ···················· | ····················· | ···················· | ···················· | ···················· |
| P | 5394 hCoV-19/Denmark/DCGC-889/2020\|EPI_ISL_614453\|2020- | ···················· | ···················· | ····················· | ···················· | ···················· | ···················· |
| P | 5395 hCoV-19/Denmark/DCGC-890/2020\|EPI_ISL_614454\|2020- | ···················· | ···················· | ····················· | ···················· | ···················· | ···················· |
| P | 5396 hCoV-19/Denmark/DCGC-891/2020\|EPI_ISL_614455\|2020- | ···················· | ···················· | ····················· | ···················· | ···················· | ···················· |
| P | 5397 hCoV-19/Denmark/DCGC-892/2020\|EPI_ISL_614456\|2020- | ········T··········· | ···················· | ···········T········· | ···················· | ···················· | ···················· |
| P | 5398 hCoV-19/Denmark/DCGC-893/2020\|EPI_ISL_614457\|2020- | ···················· | ···················· | ····················· | ···················· | ···················· | ···················· |
| P | 5399 hCoV-19/Denmark/DCGC-894/2020\|EPI_ISL_614458\|2020- | ···················· | ···················· | ····················· | ···················· | ···················· | ···················· |
| P | 5400 hCoV-19/Denmark/DCGC-895/2020\|EPI_ISL_614459\|2020- | ···················· | ···················· | ····················· | ···················· | ···················· | ···················· |

| | | | | | | | | |
|---|---|---|---|---|---|---|---|---|
| P | 5401 | hCoV-19/Denmark/DCGC-896/2020\|EPI_ISL_614460\|2020- | ···················· | ···················· | ····················· | ···················· | ···················· | ···················· |
| P | 5402 | hCoV-19/Denmark/DCGC-897/2020\|EPI_ISL_614461\|2020- | ···················· | ···················· | ····················· | ···················· | ···················· | ···················· |
| P | 5403 | hCoV-19/Denmark/DCGC-898/2020\|EPI_ISL_614462\|2020- | ···················· | ···················· | ····················· | ···················· | ···················· | ···················· |
| P | 5404 | hCoV-19/Denmark/DCGC-899/2020\|EPI_ISL_614463\|2020- | ···················· | ···················· | ····················· | ···················· | ···················· | ···················· |
| P | 5405 | hCoV-19/Denmark/DCGC-900/2020\|EPI_ISL_614464\|2020- | ···················· | ···················· | ····················· | ···················· | ···················· | ···················· |
| P | 5406 | hCoV-19/Denmark/DCGC-902/2020\|EPI_ISL_614465\|2020- | ···················· | ···················· | ····················· | ···················· | ···················· | ···················· |
| P | 5407 | hCoV-19/Denmark/DCGC-903/2020\|EPI_ISL_614466\|2020- | ···················· | ···················· | ····················· | ···················· | ···················· | ···················· |
| P | 5408 | hCoV-19/Denmark/DCGC-904/2020\|EPI_ISL_614467\|2020- | ···················· | ···················· | ····················· | ···················· | ···················· | ···················· |
| P | 5409 | hCoV-19/Denmark/DCGC-905/2020\|EPI_ISL_614468\|2020- | ···················· | ···················· | ····················· | ···················· | ···················· | ···················· |
| P | 5410 | hCoV-19/Denmark/DCGC-906/2020\|EPI_ISL_614469\|2020- | ···················· | ···················· | ····················· | ···················· | ···················· | ···················· |
| P | 5411 | hCoV-19/Denmark/DCGC-907/2020\|EPI_ISL_614470\|2020- | ···················· | ···················· | ····················· | ···················· | ···················· | ···················· |
| P | 5412 | hCoV-19/Denmark/DCGC-908/2020\|EPI_ISL_614471\|2020- | ···················· | ···················· | ····················· | ···················· | ···················· | ···················· |
| P | 5413 | hCoV-19/Denmark/DCGC-909/2020\|EPI_ISL_614472\|2020- | ···················· | ···················· | ····················· | ···················· | ···················· | ···················· |
| P | 5414 | hCoV-19/Denmark/DCGC-1012/2020\|EPI_ISL_614473\|2020 | ···················· | ···················· | ····················· | ···················· | ···················· | ···················· |
| P | 5415 | hCoV-19/Denmark/DCGC-1013/2020\|EPI_ISL_614474\|2020 | ···················· | ···················· | ····················· | ···················· | ···················· | ···················· |
| P | 5416 | hCoV-19/Denmark/DCGC-1014/2020\|EPI_ISL_614475\|2020 | ···················· | ···················· | ····················· | ···················· | ···················· | ···················· |
| P | 5417 | hCoV-19/Denmark/DCGC-1015/2020\|EPI_ISL_614476\|2020 | ···················· | ···················· | ····················· | ···················· | ···················· | ···················· |
| P | 5418 | hCoV-19/Denmark/DCGC-1016/2020\|EPI_ISL_614477\|2020 | ···················· | ···················· | ····················· | ···················· | ···················· | ···················· |
| P | 5419 | hCoV-19/Denmark/DCGC-1017/2020\|EPI_ISL_614478\|2020 | ···················· | ···················· | ····················· | ···················· | ···················· | ···················· |
| P | 5420 | hCoV-19/Denmark/DCGC-1018/2020\|EPI_ISL_614479\|2020 | ···················· | ···················· | ····················· | ···················· | ···················· | ···················· |
| P | 5421 | hCoV-19/Denmark/DCGC-1020/2020\|EPI_ISL_614480\|2020 | ···················· | ···················· | ····················· | ···················· | ···················· | ···················· |
| P | 5422 | hCoV-19/Denmark/DCGC-1021/2020\|EPI_ISL_614481\|2020 | ···················· | ···················· | ····················· | ···················· | ···················· | ···················· |
| P | 5423 | hCoV-19/Denmark/DCGC-1022/2020\|EPI_ISL_614482\|2020 | ···················· | ···················· | ····················· | ···················· | ···················· | ···················· |
| P | 5424 | hCoV-19/Denmark/DCGC-1023/2020\|EPI_ISL_614483\|2020 | ···················· | ···················· | ····················· | ···················· | ···················· | ···················· |
| P | 5425 | hCoV-19/Denmark/DCGC-1024/2020\|EPI_ISL_614484\|2020 | ···················· | ···················· | ····················· | ···················· | ···················· | ···················· |
| P | 5426 | hCoV-19/Denmark/DCGC-1025/2020\|EPI_ISL_614485\|2020 | ···················· | ···················· | ····················· | ···················· | ···················· | ···················· |
| P | 5427 | hCoV-19/Denmark/DCGC-1026/2020\|EPI_ISL_614486\|2020 | ···················· | ···················· | ····················· | ···················· | ···················· | ···················· |
| P | 5428 | hCoV-19/Denmark/DCGC-1027/2020\|EPI_ISL_614487\|2020 | ···················· | ···················· | ····················· | ···················· | ···················· | ···················· |
| P | 5429 | hCoV-19/Denmark/DCGC-1028/2020\|EPI_ISL_614488\|2020 | ···················· | ···················· | ····················· | ···················· | ···················· | ···················· |
| P | 5430 | hCoV-19/Denmark/DCGC-1064/2020\|EPI_ISL_614489\|2020 | ···················· | ···················· | ····················· | ···················· | ···················· | ···················· |
| P | 5431 | hCoV-19/Denmark/DCGC-1065/2020\|EPI_ISL_614490\|2020 | ···················· | ···················· | ····················· | ···················· | ···················· | ···················· |
| P | 5432 | hCoV-19/Denmark/DCGC-1066/2020\|EPI_ISL_614491\|2020 | ···················· | ···················· | ····················· | ···················· | ···················· | ···················· |
| P | 5433 | hCoV-19/Denmark/DCGC-1067/2020\|EPI_ISL_614492\|2020 | ···················· | ···················· | ····················· | ···················· | ···················· | ···················· |
| P | 5434 | hCoV-19/Denmark/DCGC-1068/2020\|EPI_ISL_614493\|2020 | ···················· | ···················· | ····················· | ···················· | ···················· | ···················· |
| P | 5435 | hCoV-19/Denmark/DCGC-1069/2020\|EPI_ISL_614494\|2020 | ···················· | ···················· | ····················· | ···················· | ···················· | ···················· |
| P | 5436 | hCoV-19/Denmark/DCGC-1070/2020\|EPI_ISL_614495\|2020 | ···················· | ···················· | ····················· | ···················· | ···················· | ···················· |
| P | 5437 | hCoV-19/Denmark/DCGC-1071/2020\|EPI_ISL_614496\|2020 | ···················· | ···················· | ····················· | ···················· | ···················· | ···················· |
| P | 5438 | hCoV-19/Denmark/DCGC-1072/2020\|EPI_ISL_614497\|2020 | ···················· | ···················· | ····················· | ···················· | ···················· | ···················· |
| P | 5439 | hCoV-19/Denmark/DCGC-1073/2020\|EPI_ISL_614498\|2020 | ···················· | ···················· | ····················· | ···················· | ···················· | ···················· |
| P | 5440 | hCoV-19/Denmark/DCGC-1074/2020\|EPI_ISL_614499\|2020 | ···················· | ···················· | ····················· | ···················· | ···················· | ···················· |
| P | 5441 | hCoV-19/Denmark/DCGC-4290/2020\|EPI_ISL_615727\|2020 | ···················· | ···················· | ····················· | ···················· | ···················· | ···················· |
| P | 5442 | hCoV-19/Denmark/DCGC-4291/2020\|EPI_ISL_615728\|2020 | ···················· | ···················· | ····················· | ···················· | ···················· | ···················· |
| P | 5443 | hCoV-19/Denmark/DCGC-4293/2020\|EPI_ISL_615730\|2020 | ···················· | ···················· | ····················· | ···················· | ···················· | ···················· |
| P | 5444 | hCoV-19/Denmark/DCGC-4294/2020\|EPI_ISL_615731\|2020 | ···················· | ···················· | ····················· | ···················· | ···················· | ···················· |
| P | 5445 | hCoV-19/Denmark/DCGC-4295/2020\|EPI_ISL_615732\|2020 | ···················· | ···················· | ····················· | ···················· | ···················· | ···················· |
| P | 5446 | hCoV-19/Denmark/DCGC-4296/2020\|EPI_ISL_615733\|2020 | ···················· | ···················· | ····················· | ···················· | ···················· | ···················· |
| P | 5447 | hCoV-19/Denmark/DCGC-4297/2020\|EPI_ISL_615734\|2020 | ···················· | ···················· | ····················· | ···················· | ···················· | ···················· |
| P | 5448 | hCoV-19/Denmark/DCGC-4298/2020\|EPI_ISL_615735\|2020 | ···················· | ···················· | ····················· | ···················· | ···················· | ···················· |
| P | 5449 | hCoV-19/Denmark/DCGC-4299/2020\|EPI_ISL_615736\|2020 | ···················· | ···················· | ····················· | ···················· | ···················· | ···················· |
| P | 5450 | hCoV-19/Denmark/DCGC-4300/2020\|EPI_ISL_615737\|2020 | ···················· | ···················· | ····················· | ···················· | ···················· | ···················· |
| P | 5451 | hCoV-19/Denmark/DCGC-4302/2020\|EPI_ISL_615739\|2020 | ···················· | ···················· | ····················· | ···················· | ···················· | ···················· |
| P | 5452 | hCoV-19/Denmark/DCGC-4303/2020\|EPI_ISL_615740\|2020 | ···················· | ···················· | ····················· | ···················· | ···················· | ···················· |
| P | 5453 | hCoV-19/Denmark/DCGC-4305/2020\|EPI_ISL_615742\|2020 | ···················· | ···················· | ····················· | ···················· | ···················· | ···················· |
| P | 5454 | hCoV-19/Denmark/DCGC-4307/2020\|EPI_ISL_615744\|2020 | ···················· | ···················· | ····················· | ···················· | ···················· | ···················· |
| P | 5455 | hCoV-19/Denmark/DCGC-4308/2020\|EPI_ISL_615745\|2020 | ···················· | ···················· | ····················· | ···················· | ···················· | ···················· |
| P | 5456 | hCoV-19/Denmark/DCGC-4309/2020\|EPI_ISL_615746\|2020 | ···················· | ···················· | ····················· | ···················· | ···················· | ···················· |
| P | 5457 | hCoV-19/Denmark/DCGC-4310/2020\|EPI_ISL_615747\|2020 | ···················· | ···················· | ····················· | ···················· | ···················· | ···················· |
| P | 5458 | hCoV-19/Denmark/DCGC-4311/2020\|EPI_ISL_615748\|2020 | ···················· | ···················· | ····················· | ···················· | ···················· | ···················· |
| P | 5459 | hCoV-19/Denmark/DCGC-4312/2020\|EPI_ISL_615749\|2020 | ···················· | ···················· | ····················· | ···················· | ···················· | ···················· |
| P | 5460 | hCoV-19/Denmark/DCGC-4313/2020\|EPI_ISL_615750\|2020 | ···················· | ···················· | ····················· | ···················· | ···················· | ···················· |
| P | 5461 | hCoV-19/Denmark/DCGC-4314/2020\|EPI_ISL_615751\|2020 | ···················· | ···················· | ····················· | ···················· | ···················· | ···················· |
| P | 5462 | hCoV-19/Denmark/DCGC-4315/2020\|EPI_ISL_615752\|2020 | ···················· | ···················· | ····················· | ···················· | ···················· | ···················· |
| P | 5463 | hCoV-19/Denmark/DCGC-3984/2020\|EPI_ISL_615754\|2020 | ···················· | ···················· | ····················· | ···················· | ···················· | ···················· |
| P | 5464 | hCoV-19/Denmark/DCGC-3985/2020\|EPI_ISL_615755\|2020 | ········T··········· | ···················· | ····················· | ···················· | ···················· | ···················· |
| P | 5465 | hCoV-19/Denmark/DCGC-3986/2020\|EPI_ISL_615756\|2020 | ···················· | ···················· | ····················· | ···················· | ···················· | ···················· |
| P | 5466 | hCoV-19/Denmark/DCGC-3987/2020\|EPI_ISL_615757\|2020 | ···················· | ···················· | ····················· | ···················· | ···················· | ···················· |
| P | 5467 | hCoV-19/Denmark/DCGC-3988/2020\|EPI_ISL_615758\|2020 | ···················· | ···················· | ····················· | ···················· | ···················· | ···················· |
| P | 5468 | hCoV-19/Denmark/DCGC-3989/2020\|EPI_ISL_615759\|2020 | ···················· | ···················· | ····················· | ···················· | ···················· | ···················· |
| P | 5469 | hCoV-19/Denmark/DCGC-3990/2020\|EPI_ISL_615760\|2020 | ···················· | ···················· | ····················· | ···················· | ···················· | ···················· |
| P | 5470 | hCoV-19/Denmark/DCGC-3991/2020\|EPI_ISL_615761\|2020 | ···················· | ···················· | ····················· | ···················· | ···················· | ···················· |
| P | 5471 | hCoV-19/Denmark/DCGC-3992/2020\|EPI_ISL_615762\|2020 | ···················· | ···················· | ····················· | ···················· | ···················· | ···················· |
| P | 5472 | hCoV-19/Denmark/DCGC-3993/2020\|EPI_ISL_615763\|2020 | ···················· | ···················· | ····················· | ···················· | ···················· | ···················· |
| P | 5473 | hCoV-19/Denmark/DCGC-3994/2020\|EPI_ISL_615764\|2020 | ···················· | ···················· | ····················· | ···················· | ···················· | ···················· |
| P | 5474 | hCoV-19/Denmark/DCGC-3995/2020\|EPI_ISL_615765\|2020 | ···················· | ···················· | ····················· | ···················· | ···················· | ···················· |
| P | 5475 | hCoV-19/Denmark/DCGC-3996/2020\|EPI_ISL_615766\|2020 | ···················· | ···················· | ····················· | ···················· | ···················· | ···················· |
| P | 5476 | hCoV-19/Denmark/DCGC-3997/2020\|EPI_ISL_615767\|2020 | ···················· | ···················· | ····················· | ···················· | ···················· | ···················· |
| P | 5477 | hCoV-19/Denmark/DCGC-3998/2020\|EPI_ISL_615768\|2020 | ···················· | ···················· | ····················· | ···················· | ···················· | ···················· |
| P | 5478 | hCoV-19/Denmark/DCGC-3999/2020\|EPI_ISL_615769\|2020 | ···················· | ···················· | ····················· | ···················· | ···················· | ···················· |
| P | 5479 | hCoV-19/Denmark/DCGC-4008/2020\|EPI_ISL_615780\|2020 | ···················· | ···················· | ····················· | ···················· | ···················· | ···················· |
| P | 5480 | hCoV-19/Denmark/DCGC-4009/2020\|EPI_ISL_615781\|2020 | ···················· | ···················· | ····················· | ···················· | ···················· | ···················· |

P 5481 hCoV-19/Denmark/DCGC-4010/2020|EPI_ISL_615782|2020 ···················· ···················· ····················· ···················· ···················· ····················
P 5482 hCoV-19/Denmark/DCGC-4011/2020|EPI_ISL_615783|2020 ···················· ···················· ····················· ···················· ···················· ····················
P 5483 hCoV-19/Denmark/DCGC-4012/2020|EPI_ISL_615784|2020 ···················· ···················· ····················· ···················· ···················· ····················
P 5484 hCoV-19/Denmark/DCGC-4013/2020|EPI_ISL_615785|2020 ···················· ···················· ····················· ···················· ···················· ····················
P 5485 hCoV-19/Denmark/DCGC-4014/2020|EPI_ISL_615786|2020 ···················· ···················· ····················· ···················· ···················· ····················
P 5486 hCoV-19/Denmark/DCGC-4015/2020|EPI_ISL_615787|2020 ···················· ···················· ····················· ···················· ···················· ····················
P 5487 hCoV-19/Denmark/DCGC-4016/2020|EPI_ISL_615788|2020 ···················· ···················· ····················· ···················· ···················· ····················
P 5488 hCoV-19/Denmark/DCGC-4017/2020|EPI_ISL_615789|2020 ···················· ···················· ····················· ···················· ···················· ····················
P 5489 hCoV-19/Denmark/DCGC-4018/2020|EPI_ISL_615790|2020 ···················· ···················· ····················· ···················· ···················· ····················
P 5490 hCoV-19/Denmark/DCGC-4019/2020|EPI_ISL_615791|2020 ···················· ···················· ····················· ···················· ···················· ····················
P 5491 hCoV-19/Denmark/DCGC-4020/2020|EPI_ISL_615792|2020 ···················· ···················· ····················· ···················· ···················· ····················
P 5492 hCoV-19/Denmark/DCGC-4021/2020|EPI_ISL_615793|2020 ···················· ···················· ····················· ···················· ···················· ····················
P 5493 hCoV-19/Denmark/DCGC-4022/2020|EPI_ISL_615794|2020 ···················· ···················· ····················· ···················· ···················· ····················
P 5494 hCoV-19/Denmark/DCGC-4023/2020|EPI_ISL_615795|2020 ···················· ···················· ····················· ···················· ···················· ····················
P 5495 hCoV-19/Denmark/DCGC-4024/2020|EPI_ISL_615796|2020 ···················· ···················· ····················· ···················· ···················· ····················
P 5496 hCoV-19/Denmark/DCGC-4025/2020|EPI_ISL_615797|2020 ···················· ···················· ····················· ···················· ···················· ····················
P 5497 hCoV-19/Denmark/DCGC-4026/2020|EPI_ISL_615798|2020 ···················· ···················· ····················· ···················· ···················· ····················
P 5498 hCoV-19/Denmark/DCGC-4027/2020|EPI_ISL_615799|2020 ···················· ···················· ····················· ···················· ···················· ····················
P 5499 hCoV-19/Denmark/DCGC-7220/2020|EPI_ISL_619210|2020 ···················· ···················· ····················· ···················· ···················· ····················
P 5500 hCoV-19/Denmark/DCGC-1565/2020|EPI_ISL_619211|2020 ···················· ···················· ····················· ···················· ···················· ····················
P 5501 hCoV-19/Denmark/DCGC-7224/2020|EPI_ISL_619212|2020 ···················· ···················· ····················· ···················· ···················· ····················
P 5502 hCoV-19/Denmark/DCGC-7228/2020|EPI_ISL_619213|2020 ···················· ···················· ····················· ···················· ···················· ····················
P 5503 hCoV-19/Denmark/DCGC-7229/2020|EPI_ISL_619214|2020 ···················· ···················· ····················· ···················· ···················· ····················
P 5504 hCoV-19/Denmark/DCGC-7231/2020|EPI_ISL_619215|2020 ···················· ···················· ····················· ···················· ···················· ····················
P 5505 hCoV-19/Denmark/DCGC-7233/2020|EPI_ISL_619216|2020 ···················· ···················· ····················· ···················· ···················· ····················
P 5506 hCoV-19/Denmark/DCGC-7235/2020|EPI_ISL_619217|2020 ···················· ···················· ····················· ···················· ···················· ····················
P 5507 hCoV-19/Denmark/DCGC-7236/2020|EPI_ISL_619218|2020 ···················· ···················· ····················· ···················· ···················· ····················
P 5508 hCoV-19/Denmark/DCGC-7238/2020|EPI_ISL_619219|2020 ···················· ···················· ····················· ···················· ···················· ····················
P 5509 hCoV-19/Denmark/DCGC-7252/2020|EPI_ISL_619229|2020 ···················· ···················· ····················· ···················· ···················· ····················
P 5510 hCoV-19/Denmark/DCGC-5939/2020|EPI_ISL_620204|2020 ···················· ···················· ····················· ···················· ···················· ····················
P 5511 hCoV-19/Denmark/DCGC-5940/2020|EPI_ISL_620205|2020 ···················· ···················· ····················· ···················· ···················· ····················
P 5512 hCoV-19/Denmark/DCGC-5941/2020|EPI_ISL_620206|2020 ···················· ···················· ····················· ···················· ···················· ····················
P 5513 hCoV-19/Denmark/DCGC-5942/2020|EPI_ISL_620207|2020 ···················· ···················· ····················· ···················· ···················· ····················
P 5514 hCoV-19/Denmark/DCGC-5943/2020|EPI_ISL_620208|2020 ···················· ···················· ····················· ···················· ···················· ····················
P 5515 hCoV-19/Denmark/DCGC-5944/2020|EPI_ISL_620209|2020 ···················· ···················· ····················· ···················· ···················· ····················
P 5516 hCoV-19/Denmark/DCGC-5945/2020|EPI_ISL_620210|2020 ···················· ···················· ····················· ···················· ···················· ····················
P 5517 hCoV-19/Denmark/DCGC-5946/2020|EPI_ISL_620211|2020 ···················· ···················· ····················· ···················· ···················· ····················
P 5518 hCoV-19/Denmark/DCGC-5948/2020|EPI_ISL_620213|2020 ···················· ···················· ····················· ···················· ···················· ····················
P 5519 hCoV-19/Denmark/DCGC-5949/2020|EPI_ISL_620214|2020 ···················· ···················· ····················· ···················· ···················· ····················
P 5520 hCoV-19/Denmark/DCGC-4841/2020|EPI_ISL_621526|2020 ···················· ···················· ····················· ···················· ···················· ····················
P 5521 hCoV-19/Denmark/DCGC-4842/2020|EPI_ISL_621527|2020 ···················· ···················· ····················· ···················· ···················· ····················
P 5522 hCoV-19/Denmark/DCGC-4843/2020|EPI_ISL_621528|2020 ···················· ···················· ····················· ···················· ···················· ····················
P 5523 hCoV-19/Denmark/DCGC-4844/2020|EPI_ISL_621529|2020 ···················· ···················· ····················· ···················· ···················· ····················
P 5524 hCoV-19/Denmark/DCGC-1673/2020|EPI_ISL_621535|2020 ···················· ···················· ····················· ···················· ···················· ····················
P 5525 hCoV-19/Denmark/DCGC-4850/2020|EPI_ISL_621536|2020 ···················· ···················· ····················· ···················· ···················· ····················
P 5526 hCoV-19/Denmark/DCGC-4851/2020|EPI_ISL_621537|2020 ···················· ···················· ····················· ···················· ···················· ····················
P 5527 hCoV-19/Denmark/DCGC-4852/2020|EPI_ISL_621538|2020 ···················· ···················· ····················· ···················· ···················· ····················
P 5528 hCoV-19/Denmark/DCGC-4853/2020|EPI_ISL_621539|2020 ···················· ···················· ····················· ···················· ···················· ····················
P 5529 hCoV-19/Denmark/DCGC-4854/2020|EPI_ISL_621540|2020 ···················· ···················· ····················· ···················· ···················· ····················
P 5530 hCoV-19/Denmark/DCGC-4855/2020|EPI_ISL_621541|2020 ···················· ···················· ····················· ···················· ···················· ····················
P 5531 hCoV-19/Denmark/DCGC-4856/2020|EPI_ISL_621542|2020 ···················· ···················· ····················· ···················· ···················· ····················
P 5532 hCoV-19/Denmark/DCGC-4857/2020|EPI_ISL_621543|2020 ···················· ···················· ····················· ···················· ···················· ····················
P 5533 hCoV-19/Denmark/DCGC-4859/2020|EPI_ISL_621545|2020 ···················· ···················· ····················· ···················· ···················· ····················
P 5534 hCoV-19/Denmark/DCGC-5062/2020|EPI_ISL_621600|2020 ···················· ···················· ····················· ···················· ···················· ····················
P 5535 hCoV-19/England/204290951/2020|EPI_ISL_622866|2020 ···················· ···················· ····················· ···················· ···················· ····················
P 5536 hCoV-19/England/204290952/2020|EPI_ISL_622867|2020 ···················· ···················· ····················· ···················· ···················· ····················
P 5537 hCoV-19/England/204291029/2020|EPI_ISL_622868|2020 ···················· ···················· ····················· ···················· ···················· ····················
P 5538 hCoV-19/England/204291046/2020|EPI_ISL_622869|2020 ···················· ···················· ····················· ···················· ···················· ····················
P 5539 hCoV-19/England/204291054/2020|EPI_ISL_622870|2020 ···················· ···················· ····················· ···················· ···················· ····················
P 5540 hCoV-19/England/204291428/2020|EPI_ISL_622871|2020 ···················· ···················· ····················· ···················· ···················· ····················
P 5541 hCoV-19/England/204291435/2020|EPI_ISL_622872|2020 ···················· ···················· ····················· ···················· ···················· ····················
P 5542 hCoV-19/England/204291436/2020|EPI_ISL_622873|2020 ···················· ···················· ····················· ···················· ···················· ····················
P 5543 hCoV-19/England/204291437/2020|EPI_ISL_622874|2020 ···················· ···················· ····················· ···················· ···················· ····················
P 5544 hCoV-19/England/204291438/2020|EPI_ISL_622875|2020 ···················· ···················· ····················· ···················· ···················· ····················
P 5545 hCoV-19/South Africa/R13454-20/2020|EPI_ISL_622922 ···················· ···················· ····················· ···················· ···················· ····················
P 5546 hCoV-19/England/ALDP-AA2BF6/2020|EPI_ISL_625100|20 ···················· ·A·················· ····················· ···················· ···················· ····················
P 5547 hCoV-19/England/ALDP-AA36C7/2020|EPI_ISL_625101|20 ···················· ···················· ····················· ···················· ···················· ····················
P 5548 hCoV-19/England/ALDP-AA21E5/2020|EPI_ISL_625104|20 ···················· ···················· ····················· ···················· ···················· ····················
P 5549 hCoV-19/England/ALDP-AA3F80/2020|EPI_ISL_625105|20 ···················· ···················· ····················· ···················· ···················· ····················
P 5550 hCoV-19/Wales/ALDP-AA08D3/2020|EPI_ISL_625106|2020 ···················· ···················· ····················· ···················· ···················· ····················
P 5551 hCoV-19/England/ALDP-AA3630/2020|EPI_ISL_625107|20 ···················· ···················· ····················· ···················· ···················· ····················
P 5552 hCoV-19/England/ALDP-AA175D/2020|EPI_ISL_625108|20 ···················· ···················· ····················· ···················· ···················· ····················
P 5553 hCoV-19/England/ALDP-AA092B/2020|EPI_ISL_625109|20 ···················· ···················· ····················· ···················· ···················· ····················
P 5554 hCoV-19/England/ALDP-AA119B/2020|EPI_ISL_625110|20 ···················· ···················· ····················· ···················· ···················· ····················
P 5555 hCoV-19/England/ALDP-AA3418/2020|EPI_ISL_625111|20 ·······T············ ···················· ····················· ···················· ···················· ····················
P 5556 hCoV-19/England/ALDP-AA0888/2020|EPI_ISL_625112|20 ···················· ···················· ····················· ···················· ···················· ····················
P 5557 hCoV-19/England/ALDP-AA3ECF/2020|EPI_ISL_625113|20 ···················· ···················· ····················· ···················· ···················· ····················
P 5558 hCoV-19/England/ALDP-AA5346/2020|EPI_ISL_625114|20 ···················· ···················· ····················· ···················· ···················· ····················
P 5559 hCoV-19/England/ALDP-AA0EFF/2020|EPI_ISL_625115|20 ···················· ···················· ····················· ···················· ···················· ····················
P 5560 hCoV-19/England/ALDP-AA52FE/2020|EPI_ISL_625116|20 ···················· ···················· ····················· ···················· ···················· ····················

P 5561 hCoV-19/England/ALDP-AA57D1/2020|EPI_ISL_625117|20 ···················· ·A·················· ···················· ···················· ···················· ···A················
P 5562 hCoV-19/England/ALDP-AA226A/2020|EPI_ISL_625118|20 ···················· ···················· ···················· ···················· ···················· ····················
P 5563 hCoV-19/England/ALDP-AA5285/2020|EPI_ISL_625119|20 ···················· ···················· ···················· ···················· ···················· ····················
P 5564 hCoV-19/England/ALDP-AA082E/2020|EPI_ISL_625120|20 ···················· ···················· ···················· ···················· ···················· ····················
P 5565 hCoV-19/England/ALDP-AA55F5/2020|EPI_ISL_625122|20 ···················· ···················· ···················· ···················· ···················· ····················
P 5566 hCoV-19/England/ALDP-AA2BE7/2020|EPI_ISL_625123|20 ···················· ···················· ···················· ···················· ···················· ····················
P 5567 hCoV-19/England/ALDP-AA53DD/2020|EPI_ISL_625124|20 ···················· ···················· ···················· ···················· ···················· ····················
P 5568 hCoV-19/England/ALDP-AA5531/2020|EPI_ISL_625125|20 ···················· ···················· ···················· ···················· ···················· ····················
P 5569 hCoV-19/England/ALDP-AA3F26/2020|EPI_ISL_625126|20 ···················· ·A·················· ···················· ···················· ···················· ····················
P 5570 hCoV-19/England/ALDP-AA572C/2020|EPI_ISL_625127|20 ···················· ···················· ···················· ···················· ···················· ····················
P 5571 hCoV-19/England/ALDP-AA0ADD/2020|EPI_ISL_625129|20 ···················· ···················· ···················· ···················· ···················· ····················
P 5572 hCoV-19/England/ALDP-AA0CD7/2020|EPI_ISL_625130|20 ···················· ···················· ···················· ···················· ···················· ····················
P 5573 hCoV-19/England/ALDP-AA0C7D/2020|EPI_ISL_625131|20 ···················· ···················· ···················· ···················· ···················· ····················
P 5574 hCoV-19/England/ALDP-AA2CE4/2020|EPI_ISL_625132|20 ···················· ···················· ···················· ···················· ···················· ····················
P 5575 hCoV-19/England/ALDP-AA0BDA/2020|EPI_ISL_625134|20 ···················· ···················· ···················· ···················· ···················· ····················
P 5576 hCoV-19/England/ALDP-AA0B16/2020|EPI_ISL_625135|20 ···················· ·A·················· ···················· ···················· ···················· ···A················
P 5577 hCoV-19/England/ALDP-AA17D5/2020|EPI_ISL_625136|20 ···················· ···················· ···················· ···················· ···················· ····················
P 5578 hCoV-19/England/ALDP-AA11AA/2020|EPI_ISL_625137|20 ···················· ···················· ···················· ···················· ···················· ····················
P 5579 hCoV-19/Wales/ALDP-AA5443/2020|EPI_ISL_625138|2020 ···················· ···················· ···················· ···················· ···················· ····················
P 5580 hCoV-19/England/ALDP-AA20F7/2020|EPI_ISL_625139|20 ···················· ···················· ···················· ···················· ···················· ····················
P 5581 hCoV-19/England/ALDP-AA2CA8/2020|EPI_ISL_625140|20 ···················· ···················· ···················· ···················· ···················· ····················
P 5582 hCoV-19/England/ALDP-AA1641/2020|EPI_ISL_625141|20 ···················· ···················· ···················· ···················· ···················· ····················
P 5583 hCoV-19/England/ALDP-AA0AFB/2020|EPI_ISL_625142|20 ···················· ···················· ···················· ···················· ···················· ····················
P 5584 hCoV-19/England/ALDP-AA53EC/2020|EPI_ISL_625143|20 ···················· ···················· ···················· ···················· ······T············· ····················
P 5585 hCoV-19/England/ALDP-AA0C9B/2020|EPI_ISL_625144|20 ···················· ···················· ···················· ···················· ···················· ····················
P 5586 hCoV-19/England/ALDP-AA1E2B/2020|EPI_ISL_625147|20 ···················· ···················· ···················· ···················· ···················· ····················
P 5587 hCoV-19/England/ALDP-AA2C20/2020|EPI_ISL_625148|20 ···················· ···················· ···················· ···················· ···················· ····················
P 5588 hCoV-19/England/ALDP-AA557D/2020|EPI_ISL_625149|20 ···················· ···················· ···················· ···················· ···················· ····················
P 5589 hCoV-19/England/ALDP-AA4A42/2020|EPI_ISL_625150|20 ···················· ···················· ···················· ···················· ···················· ····················
P 5590 hCoV-19/England/ALDP-AA224C/2020|EPI_ISL_625152|20 ···················· ···················· ···················· ···················· ···················· ····················
P 5591 hCoV-19/England/ALDP-AA5382/2020|EPI_ISL_625153|20 ···················· ···················· ···················· ···················· ···················· ····················
P 5592 hCoV-19/England/ALDP-AA1DF1/2020|EPI_ISL_625154|20 ···················· ···················· ···················· ···················· ···················· ····················
P 5593 hCoV-19/Wales/ALDP-AA0C13/2020|EPI_ISL_625155|2020 ···················· ·A·················· ···················· ···················· ···················· ····················
P 5594 hCoV-19/England/ALDP-AA0D10/2020|EPI_ISL_625156|20 ···················· ···················· ···················· ···················· ···················· ····················
P 5595 hCoV-19/England/ALDP-AA5328/2020|EPI_ISL_625157|20 ···················· ···················· ···················· ···················· ···················· ····················
P 5596 hCoV-19/England/ALDP-AA0EE0/2020|EPI_ISL_625158|20 ···················· ···················· ···················· ···················· ···················· ····················
P 5597 hCoV-19/England/ALDP-AA183C/2020|EPI_ISL_625159|20 ···················· ···················· ···················· ···················· ···················· ····················
P 5598 hCoV-19/Wales/ALDP-AA07F4/2020|EPI_ISL_625160|2020 ···················· ···················· ···················· ···················· ···················· ····················
P 5599 hCoV-19/England/ALDP-AA07E5/2020|EPI_ISL_625161|20 ···················· ·A·················· ···················· ···················· ···················· ····················
P 5600 hCoV-19/England/ALDP-AA32B4/2020|EPI_ISL_625162|20 ···················· ···················· ···················· ···················· ···················· ····················
P 5601 hCoV-19/England/ALDP-AA52EF/2020|EPI_ISL_625163|20 ···················· ···················· ···················· ···················· ······T············· ····················
P 5602 hCoV-19/England/ALDP-AA3F44/2020|EPI_ISL_625164|20 ···················· ···················· ···················· ···················· ···················· ····················
P 5603 hCoV-19/England/ALDP-AA2CB7/2020|EPI_ISL_625165|20 ···················· ···················· ···················· ···················· ···················· ····················
P 5604 hCoV-19/England/ALDP-AA3EDE/2020|EPI_ISL_625166|20 ···················· ···················· ···················· ···················· ···················· ····················
P 5605 hCoV-19/England/ALDP-AA3F9F/2020|EPI_ISL_625167|20 ···················· ···················· ···················· ···················· ···················· ····················
P 5606 hCoV-19/England/ALDP-AA0D5C/2020|EPI_ISL_625169|20 ···················· ···················· ···················· ···················· ···················· ····················
P 5607 hCoV-19/England/ALDP-AA225B/2020|EPI_ISL_625170|20 ···················· ···················· ···················· ···················· ···················· ····················
P 5608 hCoV-19/England/ALDP-AA1E0D/2020|EPI_ISL_625171|20 ···················· ···················· ···················· ···················· ···················· ····················
P 5609 hCoV-19/England/ALDP-AA4972/2020|EPI_ISL_625172|20 ···················· ···················· ···················· ···················· ···················· ····················
P 5610 hCoV-19/England/ALDP-AA380D/2020|EPI_ISL_625173|20 ···················· ···················· ···················· ···················· ···················· ····················
P 5611 hCoV-19/England/ALDP-AA1DD3/2020|EPI_ISL_625174|20 ···················· ··············C····· ···················· ···················· ···················· ····················
P 5612 hCoV-19/England/ALDP-AA086A/2020|EPI_ISL_625175|20 ···················· ···················· ···················· ···················· ···················· ····················
P 5613 hCoV-19/England/ALDP-AA49BE/2020|EPI_ISL_625176|20 ···················· ···················· ···················· ···················· ···················· ····················
P 5614 hCoV-19/England/ALDP-AA57C2/2020|EPI_ISL_625177|20 ···················· ···················· ···················· ···················· ···················· ····················
P 5615 hCoV-19/England/ALDP-AA4532/2020|EPI_ISL_625178|20 ···················· ···················· ···················· ···················· ···················· ····················
P 5616 hCoV-19/England/ALDP-AA181E/2020|EPI_ISL_625179|20 ···················· ···················· ···················· ···················· ···················· ····················
P 5617 hCoV-19/England/ALDP-AA3445/2020|EPI_ISL_625181|20 ···················· ···················· ···················· ···················· ···················· ····················
P 5618 hCoV-19/England/ALDP-AA52D0/2020|EPI_ISL_625183|20 ···················· ···················· ···················· ···················· ···················· ····················
P 5619 hCoV-19/England/ALDP-AA57E0/2020|EPI_ISL_625184|20 ···················· ···················· ···················· ···················· ···················· ····················
P 5620 hCoV-19/England/ALDP-AA5540/2020|EPI_ISL_625186|20 ···················· ···················· ···················· ···················· ···················· ····················
P 5621 hCoV-19/England/ALDP-AA2200/2020|EPI_ISL_625187|20 ···················· ···················· ···················· ···················· ···················· ····················
P 5622 hCoV-19/England/ALDP-AA0A55/2020|EPI_ISL_625188|20 ···················· ···················· ···················· ···················· ···················· ····················
P 5623 hCoV-19/England/ALDP-AA55E6/2020|EPI_ISL_625189|20 ···················· ···················· ···················· ···················· ···················· ····················
P 5624 hCoV-19/England/ALDP-AA4A06/2020|EPI_ISL_625190|20 ···················· ·A·················· ···················· ···················· ···················· ···A················
P 5625 hCoV-19/England/ALDP-AA090D/2020|EPI_ISL_625191|20 ···················· ···················· ···················· ···················· ···················· ····················
P 5626 hCoV-19/England/ALDP-AA456F/2020|EPI_ISL_625192|20 ···················· ···················· ···················· ···················· ···················· ····················
P 5627 hCoV-19/England/ALDP-AA1650/2020|EPI_ISL_625193|20 ···················· ···················· ···················· ···················· ···················· ····················
P 5628 hCoV-19/England/ALDP-AA0AA0/2020|EPI_ISL_625194|20 ···················· ···················· ···················· ···················· ···················· ····················
P 5629 hCoV-19/England/ALDP-AA0A91/2020|EPI_ISL_625196|20 ···················· ···················· ···················· ···················· ···················· ····················
P 5630 hCoV-19/England/ALDP-AA117D/2020|EPI_ISL_625197|20 ···················· ···················· ···················· ···················· ···················· ····················
P 5631 hCoV-19/England/ALDP-AA0BE9/2020|EPI_ISL_625198|20 ···················· ···················· ···················· ···················· ···················· ····················
P 5632 hCoV-19/England/ALDP-AA55D7/2020|EPI_ISL_625199|20 ···················· ···················· ···················· ···················· ···················· ···A················
P 5633 hCoV-19/Denmark/DCGC-4263/2020|EPI_ISL_615700|2020 ···················· ···················· ···················· ···················· ···················· ····················
P 5634 hCoV-19/Denmark/DCGC-4264/2020|EPI_ISL_615701|2020 ···················· ···················· ···················· ···················· ···················· ····················
P 5635 hCoV-19/Denmark/DCGC-4266/2020|EPI_ISL_615703|2020 ···················· ···················· ···················· ···················· ···················· ····················
P 5636 hCoV-19/Denmark/DCGC-4267/2020|EPI_ISL_615704|2020 ···················· ····G··············· ···················· ···················· ···················· ····················
P 5637 hCoV-19/Denmark/DCGC-4268/2020|EPI_ISL_615705|2020 ···················· ···················· ···················· ···················· ···················· ····················
P 5638 hCoV-19/Denmark/DCGC-4269/2020|EPI_ISL_615706|2020 ···················· ···················· ···················· ···················· ···················· ····················
P 5639 hCoV-19/Denmark/DCGC-4270/2020|EPI_ISL_615707|2020 ···················· ···················· ···················· ···················· ···················· ····················
P 5640 hCoV-19/Denmark/DCGC-4271/2020|EPI_ISL_615708|2020 ···················· ···················· ···················· ···················· ···················· ····················

| | | | | | | | |
|---|---|---|---|---|---|---|---|
| P | 5641 hCoV-19/Denmark/DCGC-4273/2020\|EPI_ISL_615710\|2020 | .................... | .................... | .................... | .................... | .................... | .................... |
| P | 5642 hCoV-19/Denmark/DCGC-4274/2020\|EPI_ISL_615711\|2020 | .................... | .................... | .................... | .................... | .................... | .................... |
| P | 5643 hCoV-19/Denmark/DCGC-4275/2020\|EPI_ISL_615712\|2020 | .................... | .................... | .................... | .................... | .................... | .................... |
| P | 5644 hCoV-19/Denmark/DCGC-4277/2020\|EPI_ISL_615714\|2020 | .................... | .................... | .................... | .................... | .................... | .................... |
| P | 5645 hCoV-19/Denmark/DCGC-4278/2020\|EPI_ISL_615715\|2020 | .................... | .................... | .................... | .................... | .................... | .................... |
| P | 5646 hCoV-19/Denmark/DCGC-4279/2020\|EPI_ISL_615716\|2020 | .................... | .................... | .................... | .................... | .................... | .................... |
| P | 5647 hCoV-19/Denmark/DCGC-4280/2020\|EPI_ISL_615717\|2020 | .................... | .................... | .................... | .................... | .................... | .................... |
| P | 5648 hCoV-19/Denmark/DCGC-4282/2020\|EPI_ISL_615719\|2020 | .................... | .................... | .................... | .................... | .................... | .................... |
| P | 5649 hCoV-19/Denmark/DCGC-4283/2020\|EPI_ISL_615720\|2020 | .................... | .................... | .................... | .................... | .................... | .................... |
| P | 5650 hCoV-19/Denmark/DCGC-4284/2020\|EPI_ISL_615721\|2020 | .................... | .................... | .................... | .................... | .................... | .................... |
| P | 5651 hCoV-19/Denmark/DCGC-4285/2020\|EPI_ISL_615722\|2020 | .................... | .................... | .................... | .................... | .................... | .................... |
| P | 5652 hCoV-19/Denmark/DCGC-4286/2020\|EPI_ISL_615723\|2020 | .................... | .................... | .................... | .................... | .................... | .................... |
| P | 5653 hCoV-19/Denmark/DCGC-4287/2020\|EPI_ISL_615724\|2020 | .................... | .A.................. | .................... | .................... | .................... | ...A................ |
| P | 5654 hCoV-19/Denmark/DCGC-4288/2020\|EPI_ISL_615725\|2020 | .................... | .A.................. | .................... | .................... | .................... | ...A................ |
| P | 5655 hCoV-19/Denmark/DCGC-7206/2020\|EPI_ISL_619200\|2020 | .................... | .................... | .................... | .................... | .................... | .................... |
| P | 5656 hCoV-19/Denmark/DCGC-7207/2020\|EPI_ISL_619201\|2020 | .................... | .................... | .................... | .................... | .................... | .................... |
| P | 5657 hCoV-19/Denmark/DCGC-7211/2020\|EPI_ISL_619202\|2020 | .................... | .................... | .................... | .................... | .................... | .................... |
| P | 5658 hCoV-19/Denmark/DCGC-7212/2020\|EPI_ISL_619203\|2020 | .................... | .................... | .................... | .................... | .................... | .................... |
| P | 5659 hCoV-19/Denmark/DCGC-7213/2020\|EPI_ISL_619204\|2020 | .................... | .................... | .................... | .................... | .................... | .................... |
| P | 5660 hCoV-19/Denmark/DCGC-7214/2020\|EPI_ISL_619205\|2020 | .................... | .................... | .................... | .................... | .................... | .................... |
| P | 5661 hCoV-19/Denmark/DCGC-7215/2020\|EPI_ISL_619206\|2020 | .................... | .................... | .................... | .................... | .................... | .................... |
| P | 5662 hCoV-19/Denmark/DCGC-7216/2020\|EPI_ISL_619207\|2020 | .................... | .................... | .................... | .................... | .................... | .................... |
| P | 5663 hCoV-19/Denmark/DCGC-7218/2020\|EPI_ISL_619208\|2020 | .................... | .................... | .................... | .................... | .................... | .................... |
| P | 5664 hCoV-19/Denmark/DCGC-7219/2020\|EPI_ISL_619209\|2020 | .................... | .................... | .................... | .................... | .................... | .................... |
| P | 5665 hCoV-19/Denmark/DCGC-7239/2020\|EPI_ISL_619220\|2020 | .................... | .................... | .................... | .................... | .................... | .................... |
| P | 5666 hCoV-19/Denmark/DCGC-7240/2020\|EPI_ISL_619221\|2020 | .................... | .................... | .................... | .................... | .................... | .................... |
| P | 5667 hCoV-19/Denmark/DCGC-7243/2020\|EPI_ISL_619222\|2020 | .................... | .................... | .................... | .................... | .................... | .................... |
| P | 5668 hCoV-19/Denmark/DCGC-7244/2020\|EPI_ISL_619223\|2020 | .................... | .................... | .................... | .................... | .................... | .................... |
| P | 5669 hCoV-19/Denmark/DCGC-7245/2020\|EPI_ISL_619224\|2020 | .................... | .................... | .................... | .................... | .................... | .................... |
| P | 5670 hCoV-19/Denmark/DCGC-7246/2020\|EPI_ISL_619225\|2020 | .................... | .................... | .................... | .................... | .................... | .................... |
| P | 5671 hCoV-19/Denmark/DCGC-1564/2020\|EPI_ISL_619226\|2020 | .................... | .................... | .................... | .................... | .................... | .................... |
| P | 5672 hCoV-19/Denmark/DCGC-7249/2020\|EPI_ISL_619227\|2020 | .................... | .................... | .................... | .................... | .................... | .................... |
| P | 5673 hCoV-19/Denmark/DCGC-7250/2020\|EPI_ISL_619228\|2020 | .................... | .................... | .................... | .................... | .................... | .................... |
| P | 5674 hCoV-19/Denmark/DCGC-7253/2020\|EPI_ISL_619230\|2020 | .................... | .................... | .................... | .................... | .................... | .................... |
| P | 5675 hCoV-19/Denmark/DCGC-7255/2020\|EPI_ISL_619231\|2020 | .................... | .................... | .................... | .................... | .................... | .................... |
| P | 5676 hCoV-19/Denmark/DCGC-7256/2020\|EPI_ISL_619232\|2020 | .................... | .................... | .................... | .................... | .................... | .................... |
| P | 5677 hCoV-19/Denmark/DCGC-1631/2020\|EPI_ISL_619233\|2020 | .................... | .................... | .................... | .................... | .................... | .................... |
| P | 5678 hCoV-19/Denmark/DCGC-7257/2020\|EPI_ISL_619234\|2020 | .................... | .................... | .................... | .................... | .................... | .................... |
| P | 5679 hCoV-19/Denmark/DCGC-7259/2020\|EPI_ISL_619235\|2020 | .................... | .A.................. | .................... | .................... | .................... | .................... |
| P | 5680 hCoV-19/Denmark/DCGC-7260/2020\|EPI_ISL_619236\|2020 | .................... | .................... | .................... | .................... | .................... | .................... |
| P | 5681 hCoV-19/Denmark/DCGC-7261/2020\|EPI_ISL_619237\|2020 | .................... | .................... | .................... | .................... | .................... | .................... |
| P | 5682 hCoV-19/Denmark/DCGC-7264/2020\|EPI_ISL_619238\|2020 | .................... | .................... | .................... | .................... | .................... | .................... |
| P | 5683 hCoV-19/Denmark/DCGC-7266/2020\|EPI_ISL_619239\|2020 | .................... | .................... | .................... | .................... | .................... | .................... |
| P | 5684 hCoV-19/Denmark/DCGC-7267/2020\|EPI_ISL_619240\|2020 | .................... | .................... | .................... | .................... | .................... | .................... |
| P | 5685 hCoV-19/Denmark/DCGC-7268/2020\|EPI_ISL_619241\|2020 | .................... | .A.................. | .................... | .................... | .................... | .................... |
| P | 5686 hCoV-19/Denmark/DCGC-7270/2020\|EPI_ISL_619242\|2020 | .................... | .................... | .................... | .................... | .................... | .................... |
| P | 5687 hCoV-19/Denmark/DCGC-7272/2020\|EPI_ISL_619243\|2020 | .................... | .................... | .................... | .................... | .................... | .................... |
| P | 5688 hCoV-19/Denmark/DCGC-7273/2020\|EPI_ISL_619244\|2020 | .................... | .................... | .................... | .................... | .................... | .................... |
| P | 5689 hCoV-19/Denmark/DCGC-7274/2020\|EPI_ISL_619245\|2020 | .................... | .................... | .................... | .................... | .................... | .................... |
| P | 5690 hCoV-19/Denmark/DCGC-7275/2020\|EPI_ISL_619246\|2020 | .................... | .................... | .................... | .................... | .................... | .................... |
| P | 5691 hCoV-19/Denmark/DCGC-7276/2020\|EPI_ISL_619247\|2020 | .................... | .................... | .................... | .................... | .................... | .................... |
| P | 5692 hCoV-19/Denmark/DCGC-7279/2020\|EPI_ISL_619248\|2020 | .................... | .A.................. | .................... | .................... | .................... | .................... |
| P | 5693 hCoV-19/Denmark/DCGC-7283/2020\|EPI_ISL_619249\|2020 | .................... | .................... | .................... | .................... | .................... | .................... |
| P | 5694 hCoV-19/Denmark/DCGC-7284/2020\|EPI_ISL_619250\|2020 | .................... | .................... | .................... | .................... | .................... | .................... |
| P | 5695 hCoV-19/Denmark/DCGC-7285/2020\|EPI_ISL_619251\|2020 | .................... | .A.................. | .................... | .................... | .................... | .................... |
| P | 5696 hCoV-19/Denmark/DCGC-7286/2020\|EPI_ISL_619252\|2020 | .................... | .................... | .................... | .................... | .................... | .................... |
| P | 5697 hCoV-19/Denmark/DCGC-7291/2020\|EPI_ISL_619253\|2020 | .................... | .................... | .................... | .................... | .................... | .................... |
| P | 5698 hCoV-19/Denmark/DCGC-7294/2020\|EPI_ISL_619254\|2020 | .................... | .................... | .................... | .................... | .................... | .................... |
| P | 5699 hCoV-19/Denmark/DCGC-7295/2020\|EPI_ISL_619255\|2020 | .................... | .................... | .................... | .................... | .................... | .................... |
| P | 5700 hCoV-19/Denmark/DCGC-1630/2020\|EPI_ISL_619256\|2020 | .................... | .................... | .................... | .................... | .................... | .................... |
| P | 5701 hCoV-19/Denmark/DCGC-7297/2020\|EPI_ISL_619257\|2020 | .................... | .................... | .................... | .................... | .................... | .................... |
| P | 5702 hCoV-19/Denmark/DCGC-7298/2020\|EPI_ISL_619258\|2020 | .................... | .................... | .................... | .................... | .................... | .................... |
| P | 5703 hCoV-19/Denmark/DCGC-7299/2020\|EPI_ISL_619259\|2020 | .................... | .................... | .................... | .................... | .................... | .................... |
| P | 5704 hCoV-19/Denmark/DCGC-7362/2020\|EPI_ISL_619290\|2020 | .................... | .................... | .................... | .................... | .................... | .................... |
| P | 5705 hCoV-19/Denmark/DCGC-7370/2020\|EPI_ISL_619291\|2020 | .................... | .................... | .................... | .................... | .................... | .................... |
| P | 5706 hCoV-19/Denmark/DCGC-1614/2020\|EPI_ISL_619292\|2020 | .................... | .................... | .................... | .................... | .................... | .................... |
| P | 5707 hCoV-19/Denmark/DCGC-7301/2020\|EPI_ISL_619293\|2020 | .................... | .................... | .................... | .................... | .................... | .................... |
| P | 5708 hCoV-19/Denmark/DCGC-7309/2020\|EPI_ISL_619294\|2020 | .................... | .................... | .................... | .................... | .................... | .................... |
| P | 5709 hCoV-19/Denmark/DCGC-7322/2020\|EPI_ISL_619295\|2020 | .................... | .................... | .................... | .................... | .................... | .................... |
| P | 5710 hCoV-19/Denmark/DCGC-7329/2020\|EPI_ISL_619296\|2020 | .................... | .A.................. | .................... | .................... | .................... | .................... |
| P | 5711 hCoV-19/Denmark/DCGC-7335/2020\|EPI_ISL_619297\|2020 | .................... | .................... | .................... | .................... | .................... | .................... |
| P | 5712 hCoV-19/Denmark/DCGC-7343/2020\|EPI_ISL_619298\|2020 | .................... | .................... | .................... | .................... | .................... | .................... |
| P | 5713 hCoV-19/Denmark/DCGC-7378/2020\|EPI_ISL_619299\|2020 | .................... | .................... | .................... | .................... | .................... | .................... |
| P | 5714 hCoV-19/Denmark/DCGC-5935/2020\|EPI_ISL_620200\|2020 | .................... | .................... | .................... | .................... | .................... | .................... |
| P | 5715 hCoV-19/Denmark/DCGC-5936/2020\|EPI_ISL_620201\|2020 | .................... | .................... | .................... | .................... | .................... | .................... |
| P | 5716 hCoV-19/Denmark/DCGC-5937/2020\|EPI_ISL_620202\|2020 | .................... | .................... | .................... | .................... | .................... | .................... |
| P | 5717 hCoV-19/Denmark/DCGC-5938/2020\|EPI_ISL_620203\|2020 | .................... | .................... | .................... | .................... | .................... | .................... |
| P | 5718 hCoV-19/Denmark/DCGC-5950/2020\|EPI_ISL_620215\|2020 | .................... | .................... | .................... | .................... | .................... | .................... |
| P | 5719 hCoV-19/Denmark/DCGC-5951/2020\|EPI_ISL_620216\|2020 | .................... | .................... | .................... | .................... | .................... | .................... |
| P | 5720 hCoV-19/Denmark/DCGC-5952/2020\|EPI_ISL_620217\|2020 | .................... | .................... | .................... | .................... | .................... | .................... |

| | | | | | | | | |
|---|---|---|---|---|---|---|---|---|
| P | 5721 | hCoV-19/Denmark/DCGC-5953/2020\|EPI_ISL_620218\|2020 | ···················· | ···················· | ···················· | ···················· | ···················· | ···················· |
| P | 5722 | hCoV-19/Denmark/DCGC-5954/2020\|EPI_ISL_620219\|2020 | ···················· | ···················· | ···················· | ···················· | ···················· | ···················· |
| P | 5723 | hCoV-19/Denmark/DCGC-5955/2020\|EPI_ISL_620220\|2020 | ···················· | ···················· | ···················· | ···················· | ···················· | ···················· |
| P | 5724 | hCoV-19/Denmark/DCGC-5956/2020\|EPI_ISL_620221\|2020 | ···················· | ···················· | ···················· | ···················· | ······T············· | ···················· |
| P | 5725 | hCoV-19/Denmark/DCGC-5957/2020\|EPI_ISL_620222\|2020 | ···················· | ···················· | ···················· | ···················· | ···················· | ···················· |
| P | 5726 | hCoV-19/Denmark/DCGC-5958/2020\|EPI_ISL_620223\|2020 | ···················· | ···················· | ···················· | ···················· | ···················· | ···················· |
| P | 5727 | hCoV-19/Denmark/DCGC-5959/2020\|EPI_ISL_620224\|2020 | ···················· | ···················· | ···················· | ···················· | ···················· | ···················· |
| P | 5728 | hCoV-19/Denmark/DCGC-5960/2020\|EPI_ISL_620225\|2020 | ···················· | ···················· | ···················· | ···················· | ···················· | ···················· |
| P | 5729 | hCoV-19/Denmark/DCGC-6032/2020\|EPI_ISL_620226\|2020 | ···················· | ···················· | ···················· | ···················· | ···················· | ···················· |
| P | 5730 | hCoV-19/Denmark/DCGC-6033/2020\|EPI_ISL_620227\|2020 | ···················· | ···················· | ···················· | ···················· | ···················· | ···················· |
| P | 5731 | hCoV-19/Denmark/DCGC-6034/2020\|EPI_ISL_620228\|2020 | ···················· | ···················· | ···················· | ···················· | ···················· | ···················· |
| P | 5732 | hCoV-19/Denmark/DCGC-6035/2020\|EPI_ISL_620229\|2020 | ···················· | ···················· | ···················· | ···················· | ···················· | ···················· |
| P | 5733 | hCoV-19/Denmark/DCGC-6037/2020\|EPI_ISL_620231\|2020 | ···················· | ·A·················· | ···················· | ···················· | ···················· | ···················· |
| P | 5734 | hCoV-19/Denmark/DCGC-6042/2020\|EPI_ISL_620236\|2020 | ···················· | ···················· | ···················· | ···················· | ···················· | ···················· |
| P | 5735 | hCoV-19/Denmark/DCGC-6054/2020\|EPI_ISL_620248\|2020 | ···················· | ···················· | ···················· | ···················· | ···················· | ···················· |
| P | 5736 | hCoV-19/Denmark/DCGC-6055/2020\|EPI_ISL_620249\|2020 | ···················· | ···················· | ···················· | ···················· | ···················· | ···················· |
| P | 5737 | hCoV-19/Denmark/DCGC-6056/2020\|EPI_ISL_620250\|2020 | ···················· | ···················· | ···················· | ···················· | ···················· | ···················· |
| P | 5738 | hCoV-19/Denmark/DCGC-6064/2020\|EPI_ISL_620258\|2020 | ···················· | ···················· | ···················· | ···················· | ···················· | ···················· |
| P | 5739 | hCoV-19/Denmark/DCGC-6086/2020\|EPI_ISL_620280\|2020 | ···················· | ···················· | ···················· | ···················· | ···················· | ···················· |
| P | 5740 | hCoV-19/Denmark/DCGC-6087/2020\|EPI_ISL_620281\|2020 | ···················· | ···················· | ···················· | ···················· | ···················· | ···················· |
| P | 5741 | hCoV-19/Denmark/DCGC-6088/2020\|EPI_ISL_620282\|2020 | ···················· | ···················· | ···················· | ···················· | ···················· | ···················· |
| P | 5742 | hCoV-19/Denmark/DCGC-6089/2020\|EPI_ISL_620283\|2020 | ···················· | ···················· | ···················· | ···················· | ···················· | ···················· |
| P | 5743 | hCoV-19/Denmark/DCGC-6090/2020\|EPI_ISL_620284\|2020 | ···················· | ···················· | ···················· | ···················· | ···················· | ···················· |
| P | 5744 | hCoV-19/Denmark/DCGC-1283/2020\|EPI_ISL_620286\|2020 | ···················· | ···················· | ···················· | ···················· | ···················· | ···················· |
| P | 5745 | hCoV-19/Denmark/DCGC-6092/2020\|EPI_ISL_620287\|2020 | ···················· | ···················· | ···················· | ···················· | ···················· | ···················· |
| P | 5746 | hCoV-19/Denmark/DCGC-6093/2020\|EPI_ISL_620288\|2020 | ···················· | ···················· | ···················· | ···················· | ···················· | ···················· |
| P | 5747 | hCoV-19/Denmark/DCGC-6094/2020\|EPI_ISL_620289\|2020 | ···················· | ···················· | ···················· | ···················· | ···················· | ···················· |
| P | 5748 | hCoV-19/Denmark/DCGC-6095/2020\|EPI_ISL_620290\|2020 | ···················· | ···················· | ···················· | ···················· | ···················· | ···················· |
| P | 5749 | hCoV-19/Denmark/DCGC-6097/2020\|EPI_ISL_620292\|2020 | ···················· | ···················· | ···················· | ···················· | ···················· | ···················· |
| P | 5750 | hCoV-19/Denmark/DCGC-6098/2020\|EPI_ISL_620293\|2020 | ···················· | ···················· | ···················· | ···················· | ···················· | ···················· |
| P | 5751 | hCoV-19/Denmark/DCGC-6099/2020\|EPI_ISL_620294\|2020 | ···················· | ···················· | ···················· | ···················· | ···················· | ···················· |
| P | 5752 | hCoV-19/Denmark/DCGC-6100/2020\|EPI_ISL_620295\|2020 | ···················· | ···················· | ···················· | ···················· | ···················· | ···················· |
| P | 5753 | hCoV-19/Denmark/DCGC-6101/2020\|EPI_ISL_620296\|2020 | ···················· | ···················· | ···················· | ···················· | ···················· | ···················· |
| P | 5754 | hCoV-19/Denmark/DCGC-6103/2020\|EPI_ISL_620298\|2020 | ···················· | ···················· | ···················· | ···················· | ···················· | ···················· |
| P | 5755 | hCoV-19/Denmark/DCGC-6104/2020\|EPI_ISL_620299\|2020 | ···················· | ···················· | ···················· | ···················· | ···················· | ···················· |
| P | 5756 | hCoV-19/Denmark/DCGC-4962/2020\|EPI_ISL_621500\|2020 | ···················· | ···················· | ···················· | ···················· | ···················· | ···················· |
| P | 5757 | hCoV-19/Denmark/DCGC-4963/2020\|EPI_ISL_621501\|2020 | ···················· | ···················· | ···················· | ···················· | ···················· | ···················· |
| P | 5758 | hCoV-19/Denmark/DCGC-4964/2020\|EPI_ISL_621502\|2020 | ···················· | ···················· | ···················· | ···················· | ···················· | ···················· |
| P | 5759 | hCoV-19/Denmark/DCGC-4965/2020\|EPI_ISL_621503\|2020 | ···················· | ···················· | ···················· | ···················· | ···················· | ···················· |
| P | 5760 | hCoV-19/Denmark/DCGC-4966/2020\|EPI_ISL_621504\|2020 | ···················· | ···················· | ···················· | ···················· | ···················· | ···················· |
| P | 5761 | hCoV-19/Denmark/DCGC-4967/2020\|EPI_ISL_621505\|2020 | ···················· | ···················· | ···················· | ···················· | ···················· | ···················· |
| P | 5762 | hCoV-19/Denmark/DCGC-4968/2020\|EPI_ISL_621506\|2020 | ···················· | ···················· | ···················· | ···················· | ···················· | ···················· |
| P | 5763 | hCoV-19/Denmark/DCGC-4969/2020\|EPI_ISL_621507\|2020 | ···················· | ···················· | ···················· | ···················· | ···················· | ···················· |
| P | 5764 | hCoV-19/Denmark/DCGC-4970/2020\|EPI_ISL_621508\|2020 | ···················· | ···················· | ···················· | ···················· | ···················· | ···················· |
| P | 5765 | hCoV-19/Denmark/DCGC-4971/2020\|EPI_ISL_621509\|2020 | ···················· | ···················· | ···················· | ···················· | ···················· | ···················· |
| P | 5766 | hCoV-19/Denmark/DCGC-4972/2020\|EPI_ISL_621510\|2020 | ···················· | ···················· | ···················· | ···················· | ···················· | ···················· |
| P | 5767 | hCoV-19/Denmark/DCGC-4973/2020\|EPI_ISL_621511\|2020 | ···················· | ···················· | ···················· | ···················· | ···················· | ···················· |
| P | 5768 | hCoV-19/Denmark/DCGC-4974/2020\|EPI_ISL_621512\|2020 | ···················· | ···················· | ···················· | ···················· | ···················· | ···················· |
| P | 5769 | hCoV-19/Denmark/DCGC-4975/2020\|EPI_ISL_621513\|2020 | ···················· | ···················· | ···················· | ···················· | ···················· | ···················· |
| P | 5770 | hCoV-19/Denmark/DCGC-4976/2020\|EPI_ISL_621514\|2020 | ···················· | ···················· | ···················· | ···················· | ···················· | ···················· |
| P | 5771 | hCoV-19/Denmark/DCGC-4830/2020\|EPI_ISL_621515\|2020 | ···················· | ···················· | ···················· | ···················· | ···················· | ···················· |
| P | 5772 | hCoV-19/Denmark/DCGC-4831/2020\|EPI_ISL_621516\|2020 | ···················· | ···················· | ···················· | ···················· | ···················· | ···················· |
| P | 5773 | hCoV-19/Denmark/DCGC-4832/2020\|EPI_ISL_621517\|2020 | ···················· | ···················· | ···················· | ···················· | ···················· | ···················· |
| P | 5774 | hCoV-19/Denmark/DCGC-4833/2020\|EPI_ISL_621518\|2020 | ···················· | ···················· | ···················· | ···················· | ···················· | ···················· |
| P | 5775 | hCoV-19/Denmark/DCGC-4834/2020\|EPI_ISL_621519\|2020 | ···················· | ···················· | ···················· | ···················· | ···················· | ···················· |
| P | 5776 | hCoV-19/Denmark/DCGC-4835/2020\|EPI_ISL_621520\|2020 | ···················· | ···················· | ···················· | ···················· | ···················· | ···················· |
| P | 5777 | hCoV-19/Denmark/DCGC-4836/2020\|EPI_ISL_621521\|2020 | ···················· | ···················· | ···················· | ···················· | ···················· | ···················· |
| P | 5778 | hCoV-19/Denmark/DCGC-4837/2020\|EPI_ISL_621522\|2020 | ···················· | ···················· | ···················· | ···················· | ···················· | ···················· |
| P | 5779 | hCoV-19/Denmark/DCGC-4838/2020\|EPI_ISL_621523\|2020 | ···················· | ···················· | ···················· | ···················· | ···················· | ···················· |
| P | 5780 | hCoV-19/Denmark/DCGC-4839/2020\|EPI_ISL_621524\|2020 | ···················· | ···················· | ···················· | ···················· | ···················· | ···················· |
| P | 5781 | hCoV-19/Denmark/DCGC-4840/2020\|EPI_ISL_621525\|2020 | ···················· | ···················· | ···················· | ···················· | ···················· | ···················· |
| P | 5782 | hCoV-19/Denmark/DCGC-4845/2020\|EPI_ISL_621530\|2020 | ···················· | ···················· | ···················· | ···················· | ···················· | ···················· |
| P | 5783 | hCoV-19/Denmark/DCGC-4846/2020\|EPI_ISL_621531\|2020 | ···················· | ···················· | ···················· | ···················· | ···················· | ···················· |
| P | 5784 | hCoV-19/Denmark/DCGC-4847/2020\|EPI_ISL_621532\|2020 | ···················· | ···················· | ···················· | ···················· | ···················· | ···················· |
| P | 5785 | hCoV-19/Denmark/DCGC-4860/2020\|EPI_ISL_621546\|2020 | ···················· | ···················· | ···················· | ···················· | ···················· | ···················· |
| P | 5786 | hCoV-19/Denmark/DCGC-4861/2020\|EPI_ISL_621547\|2020 | ···················· | ···················· | ···················· | ···················· | ···················· | ···················· |
| P | 5787 | hCoV-19/Denmark/DCGC-4862/2020\|EPI_ISL_621548\|2020 | ···················· | ···················· | ···················· | ···················· | ···················· | ···················· |
| P | 5788 | hCoV-19/Denmark/DCGC-4863/2020\|EPI_ISL_621549\|2020 | ···················· | ···················· | ···················· | ···················· | ···················· | ···················· |
| P | 5789 | hCoV-19/Denmark/DCGC-4864/2020\|EPI_ISL_621550\|2020 | ···················· | ···················· | ···················· | ···················· | ···················· | ···················· |
| P | 5790 | hCoV-19/Denmark/DCGC-4865/2020\|EPI_ISL_621551\|2020 | ···················· | ···················· | ···················· | ···················· | ···················· | ···················· |
| P | 5791 | hCoV-19/Denmark/DCGC-4866/2020\|EPI_ISL_621552\|2020 | ···················· | ···················· | ···················· | ···················· | ···················· | ···················· |
| P | 5792 | hCoV-19/Denmark/DCGC-4867/2020\|EPI_ISL_621553\|2020 | ···················· | ···················· | ···················· | ···················· | ···················· | ···················· |
| P | 5793 | hCoV-19/Denmark/DCGC-4868/2020\|EPI_ISL_621554\|2020 | ···················· | ···················· | ···················· | ···················· | ···················· | ···················· |
| P | 5794 | hCoV-19/Denmark/DCGC-4869/2020\|EPI_ISL_621555\|2020 | ···················· | ···················· | ···················· | ···················· | ···················· | ···················· |
| P | 5795 | hCoV-19/Denmark/DCGC-4870/2020\|EPI_ISL_621556\|2020 | ···················· | ···················· | ···················· | ···················· | ···················· | ···················· |
| P | 5796 | hCoV-19/Denmark/DCGC-4871/2020\|EPI_ISL_621557\|2020 | ···················· | ···················· | ···················· | ···················· | ···················· | ···················· |
| P | 5797 | hCoV-19/Denmark/DCGC-4872/2020\|EPI_ISL_621558\|2020 | ···················· | ···················· | ···················· | ···················· | ···················· | ···················· |
| P | 5798 | hCoV-19/Denmark/DCGC-4873/2020\|EPI_ISL_621559\|2020 | ···················· | ···················· | ···················· | ···················· | ···················· | ···················· |
| P | 5799 | hCoV-19/Denmark/DCGC-4874/2020\|EPI_ISL_621560\|2020 | ···················· | ···················· | ···················· | ···················· | ···················· | ···················· |
| P | 5800 | hCoV-19/Denmark/DCGC-4876/2020\|EPI_ISL_621562\|2020 | ···················· | ···················· | ···················· | ···················· | ···················· | ···················· |

| | | | | | | | | |
|---|---|---|---|---|---|---|---|---|
| P | 5801 | hCoV-19/Denmark/DCGC-4877/2020\|EPI_ISL_621563\|2020 | ···················· | ···················· | ···················· | ···················· | ···················· | ···················· |
| P | 5802 | hCoV-19/Denmark/DCGC-4878/2020\|EPI_ISL_621564\|2020 | ···················· | ···················· | ···················· | ···················· | ···················· | ···················· |
| P | 5803 | hCoV-19/Denmark/DCGC-4879/2020\|EPI_ISL_621565\|2020 | ···················· | ···················· | ···················· | ···················· | ···················· | ···················· |
| P | 5804 | hCoV-19/Denmark/DCGC-4880/2020\|EPI_ISL_621566\|2020 | ···················· | ···················· | ···················· | ···················· | ···················· | ···················· |
| P | 5805 | hCoV-19/Denmark/DCGC-4881/2020\|EPI_ISL_621567\|2020 | ···················· | ···················· | ···················· | ···················· | ···················· | ···················· |
| P | 5806 | hCoV-19/Denmark/DCGC-4893/2020\|EPI_ISL_621579\|2020 | ···················· | ···················· | ···················· | ···················· | ···················· | ···················· |
| P | 5807 | hCoV-19/Denmark/DCGC-4894/2020\|EPI_ISL_621580\|2020 | ···················· | ···················· | ···················· | ···················· | ···················· | ···················· |
| P | 5808 | hCoV-19/Denmark/DCGC-4895/2020\|EPI_ISL_621581\|2020 | ···················· | ···················· | ···················· | ···················· | ···················· | ···················· |
| P | 5809 | hCoV-19/Denmark/DCGC-5050/2020\|EPI_ISL_621588\|2020 | ···················· | ···················· | ···················· | ···················· | ···················· | ···················· |
| P | 5810 | hCoV-19/Denmark/DCGC-5051/2020\|EPI_ISL_621589\|2020 | ···················· | ···················· | ···················· | ···················· | ···················· | ···················· |
| P | 5811 | hCoV-19/New Zealand/20CV0473/2020\|EPI_ISL_622811\|2 | ···················· | ···················· | ···················· | ···················· | ···················· | ···················· |
| P | 5812 | hCoV-19/New Zealand/20CV0474/2020\|EPI_ISL_622812\|2 | ···················· | ···················· | ···················· | ···················· | ···················· | ···················· |
| P | 5813 | hCoV-19/New Zealand/20CV0476/2020\|EPI_ISL_622813\|2 | ···················· | ···················· | ···················· | ···················· | ···················· | ···················· |
| P | 5814 | hCoV-19/New Zealand/20CV0479/2020\|EPI_ISL_622814\|2 | ···················· | ···················· | ···················· | ···················· | ···················· | ···················· |
| P | 5815 | hCoV-19/New Zealand/20CV0480/2020\|EPI_ISL_622815\|2 | ···················· | ···················· | ···················· | ···················· | ···················· | ···················· |
| P | 5816 | hCoV-19/New Zealand/20CV0481/2020\|EPI_ISL_622816\|2 | ···················· | ···················· | ···················· | ···················· | ···················· | ···················· |
| P | 5817 | hCoV-19/New Zealand/20CV0484/2020\|EPI_ISL_622817\|2 | ···················· | ···················· | ···················· | ···················· | ···················· | ···················· |
| P | 5818 | hCoV-19/New Zealand/20CV0486/2020\|EPI_ISL_622818\|2 | ···················· | ···················· | ···················· | ···················· | ···················· | ···················· |
| P | 5819 | hCoV-19/New Zealand/20CV0487/2020\|EPI_ISL_622819\|2 | ···················· | ···················· | ···················· | ···················· | ···················· | ···················· |
| P | 5820 | hCoV-19/New Zealand/20CV0488/2020\|EPI_ISL_622820\|2 | ···················· | ···················· | ···················· | ···················· | ···················· | ···················· |
| P | 5821 | hCoV-19/New Zealand/20CV0489/2020\|EPI_ISL_622821\|2 | ···················· | ·A·················· | ···················· | ···················· | ···················· | ··A················· |
| P | 5822 | hCoV-19/New Zealand/20CV0490/2020\|EPI_ISL_622822\|2 | ···················· | ·A·················· | ···················· | ···················· | ···················· | ··A················· |
| P | 5823 | hCoV-19/New Zealand/20CV0493/2020\|EPI_ISL_622823\|2 | ···················· | ···················· | ···················· | ···················· | ···················· | ···················· |
| P | 5824 | hCoV-19/New Zealand/20CV0496/2020\|EPI_ISL_622824\|2 | ···················· | ···················· | ···················· | ···················· | ···················· | ···················· |
| P | 5825 | hCoV-19/New Zealand/20CV0497/2020\|EPI_ISL_622825\|2 | ···················· | ···················· | ···················· | ···················· | ···················· | ···················· |
| P | 5826 | hCoV-19/New Zealand/20CV0498/2020\|EPI_ISL_622826\|2 | ···················· | ···················· | ···················· | ···················· | ···················· | ···················· |
| P | 5827 | hCoV-19/New Zealand/20CV0499/2020\|EPI_ISL_622827\|2 | ···················· | ···················· | ···················· | ···················· | ···················· | ···················· |
| P | 5828 | hCoV-19/New Zealand/20CV0502/2020\|EPI_ISL_622828\|2 | ···················· | ···················· | ···················· | ···················· | ···················· | ···················· |
| P | 5829 | hCoV-19/New Zealand/20CV0503/2020\|EPI_ISL_622829\|2 | ···················· | ···················· | ···················· | ···················· | ···················· | ···················· |
| P | 5830 | hCoV-19/New Zealand/20CV0504/2020\|EPI_ISL_622830\|2 | ···················· | ···················· | ···················· | ···················· | ···················· | ···················· |
| P | 5831 | hCoV-19/New Zealand/20CV0508/2020\|EPI_ISL_622831\|2 | ···················· | ···················· | ···················· | ···················· | ···················· | ···················· |
| P | 5832 | hCoV-19/New Zealand/20CV0511/2020\|EPI_ISL_622832\|2 | ···················· | ···················· | ···················· | ···················· | ···················· | ···················· |
| P | 5833 | hCoV-19/England/204040288/2020\|EPI_ISL_622833\|2020 | ···················· | ·A·················· | ···················· | ···················· | ···················· | ··A················· |
| P | 5834 | hCoV-19/England/204040292/2020\|EPI_ISL_622835\|2020 | ···················· | ·A·················· | ···················· | ···················· | ···················· | ··A················· |
| P | 5835 | hCoV-19/England/204040298/2020\|EPI_ISL_622836\|2020 | ···················· | ·A·················· | ···················· | ···················· | ···················· | ··A················· |
| P | 5836 | hCoV-19/England/204040300/2020\|EPI_ISL_622837\|2020 | ···················· | ·A·················· | ···················· | ···················· | ···················· | ··A················· |
| P | 5837 | hCoV-19/England/204040323/2020\|EPI_ISL_622838\|2020 | ···················· | ·A·················· | ···················· | ···················· | ···················· | ··A················· |
| P | 5838 | hCoV-19/England/204040324/2020\|EPI_ISL_622839\|2020 | ···················· | ·A·················· | ···················· | ···················· | ···················· | ··A················· |
| P | 5839 | hCoV-19/England/204040327/2020\|EPI_ISL_622840\|2020 | ···················· | ·A·················· | ···················· | ···················· | ···················· | ··A················· |
| P | 5840 | hCoV-19/England/204040334/2020\|EPI_ISL_622841\|2020 | ···················· | ·A·················· | ···················· | ···················· | ···················· | ··A················· |
| P | 5841 | hCoV-19/England/204040338/2020\|EPI_ISL_622842\|2020 | ···················· | ·A·················· | ···················· | ···················· | ···················· | ··A················· |
| P | 5842 | hCoV-19/England/204190425/2020\|EPI_ISL_622843\|2020 | ···················· | ···················· | ···················· | ···················· | ···················· | ···················· |
| P | 5843 | hCoV-19/England/204221331/2020\|EPI_ISL_622844\|2020 | ···················· | ···················· | ···················· | ···················· | ···················· | ···················· |
| P | 5844 | hCoV-19/England/204221332/2020\|EPI_ISL_622845\|2020 | ···················· | ···················· | ···················· | ···················· | ···················· | ···················· |
| P | 5845 | hCoV-19/England/204221335/2020\|EPI_ISL_622846\|2020 | ···················· | ···················· | ···················· | ···················· | ···················· | ···················· |
| P | 5846 | hCoV-19/England/204221337/2020\|EPI_ISL_622847\|2020 | ···················· | ···················· | ···················· | ···················· | ···················· | ···················· |
| P | 5847 | hCoV-19/England/204240972/2020\|EPI_ISL_622848\|2020 | ···················· | ···················· | ···················· | ···················· | ···················· | ···················· |
| P | 5848 | hCoV-19/England/204240973/2020\|EPI_ISL_622849\|2020 | ···················· | ·A·················· | ···················· | ···················· | ···················· | ··A················· |
| P | 5849 | hCoV-19/England/204240978/2020\|EPI_ISL_622850\|2020 | ···················· | ···················· | ···················· | ···················· | ···················· | ···················· |
| P | 5850 | hCoV-19/England/204240984/2020\|EPI_ISL_622851\|2020 | ···················· | ···················· | ···················· | ···················· | ···················· | ···················· |
| P | 5851 | hCoV-19/England/204241454/2020\|EPI_ISL_622852\|2020 | ···················· | ···················· | ···················· | ···················· | ···················· | ···················· |
| P | 5852 | hCoV-19/England/204241456/2020\|EPI_ISL_622853\|2020 | ···················· | ···················· | ···················· | ···················· | ···················· | ···················· |
| P | 5853 | hCoV-19/England/204241459/2020\|EPI_ISL_622854\|2020 | ···················· | ···················· | ···················· | ···················· | ···················· | ···················· |
| P | 5854 | hCoV-19/England/204241470/2020\|EPI_ISL_622855\|2020 | ···················· | ·A·················· | ···················· | ···················· | ···················· | ··A················· |
| P | 5855 | hCoV-19/England/204261094/2020\|EPI_ISL_622856\|2020 | ···················· | ···················· | ···················· | ···················· | ···················· | ···················· |
| P | 5856 | hCoV-19/England/204281582/2020\|EPI_ISL_622857\|2020 | ···················· | ···················· | ···················· | ···················· | ···················· | ···················· |
| P | 5857 | hCoV-19/England/204281585/2020\|EPI_ISL_622858\|2020 | ···················· | ···················· | ···················· | ···················· | ···················· | ···················· |
| P | 5858 | hCoV-19/England/204281821/2020\|EPI_ISL_622859\|2020 | ···················· | ···················· | ···················· | ···················· | ···················· | ···················· |
| P | 5859 | hCoV-19/England/204290478/2020\|EPI_ISL_622860\|2020 | ···················· | ···················· | ···················· | ···················· | ···················· | ···················· |
| P | 5860 | hCoV-19/England/204290479/2020\|EPI_ISL_622861\|2020 | ···················· | ···················· | ···················· | ···················· | ···················· | ···················· |
| P | 5861 | hCoV-19/England/204290933/2020\|EPI_ISL_622862\|2020 | ···················· | ···················· | ···················· | ···················· | ···················· | ···················· |
| P | 5862 | hCoV-19/England/204290946/2020\|EPI_ISL_622863\|2020 | ···················· | ···················· | ···················· | ···················· | ···················· | ···················· |
| P | 5863 | hCoV-19/England/204290949/2020\|EPI_ISL_622864\|2020 | ···················· | ···················· | ···················· | ···················· | ···················· | ···················· |
| P | 5864 | hCoV-19/England/204290950/2020\|EPI_ISL_622865\|2020 | ···················· | ···················· | ···················· | ···················· | ···················· | ···················· |
| P | 5865 | hCoV-19/England/204291472/2020\|EPI_ISL_622877\|2020 | ···················· | ···················· | ···················· | ···················· | ···················· | ···················· |
| P | 5866 | hCoV-19/England/204441048/2020\|EPI_ISL_622878\|2020 | ···················· | ···················· | ···················· | ···················· | ···················· | ···················· |
| P | 5867 | hCoV-19/England/204441525/2020\|EPI_ISL_622881\|2020 | ···················· | ···················· | ···················· | ···················· | ···················· | ···················· |
| P | 5868 | hCoV-19/England/204441526/2020\|EPI_ISL_622882\|2020 | ···················· | ···················· | ···················· | ···················· | ···················· | ···················· |
| P | 5869 | hCoV-19/England/204441527/2020\|EPI_ISL_622883\|2020 | ···················· | ···················· | ···················· | ···················· | ···················· | ···················· |
| P | 5870 | hCoV-19/England/204441528/2020\|EPI_ISL_622884\|2020 | ···················· | ···················· | ···················· | ···················· | ···················· | ···················· |
| P | 5871 | hCoV-19/England/204441529/2020\|EPI_ISL_622885\|2020 | ···················· | ···················· | ···················· | ···················· | ···················· | ···················· |
| P | 5872 | hCoV-19/England/204441530/2020\|EPI_ISL_622886\|2020 | ···················· | ···················· | ···················· | ···················· | ···················· | ···················· |
| P | 5873 | hCoV-19/England/204441531/2020\|EPI_ISL_622887\|2020 | ···················· | ···················· | ···················· | ···················· | ···················· | ···················· |
| P | 5874 | hCoV-19/England/204441532/2020\|EPI_ISL_622888\|2020 | ···················· | ···················· | ···················· | ···················· | ···················· | ···················· |
| P | 5875 | hCoV-19/England/204441533/2020\|EPI_ISL_622889\|2020 | ···················· | ···················· | ···················· | ···················· | ···················· | ···················· |
| P | 5876 | hCoV-19/England/204441534/2020\|EPI_ISL_622890\|2020 | ···················· | ···················· | ···················· | ···················· | ···················· | ···················· |
| P | 5877 | hCoV-19/England/204441569/2020\|EPI_ISL_622892\|2020 | ···················· | ·A·················· | ···················· | ···················· | ···················· | ···················· |
| P | 5878 | hCoV-19/England/ALDP-AA0879/2020\|EPI_ISL_625090\|20 | ···················· | ···················· | ···················· | ···················· | ···················· | ···················· |
| P | 5879 | hCoV-19/England/ALDP-AA4A8E/2020\|EPI_ISL_625091\|20 | ···················· | ···················· | ···················· | ···················· | ···················· | ···················· |
| P | 5880 | hCoV-19/England/ALDP-AA571D/2020\|EPI_ISL_625093\|20 | ···················· | ···················· | ···················· | ···················· | ···················· | ···················· |

```
P 5881 hCoV-19/England/ALDP-AA5513/2020|EPI_ISL_625094|20 ···················· ···················· ····················· ···················· ···················· ····················
P 5882 hCoV-19/England/ALDP-AA116E/2020|EPI_ISL_625095|20 ···················· ···················· ····················· ···················· ···················· ····················
P 5883 hCoV-19/England/ALDP-AA2C11/2020|EPI_ISL_625096|20 ···················· ···················· ····················· ···················· ···················· ····················
P 5884 hCoV-19/England/ALDP-AA564D/2020|EPI_ISL_625097|20 ···················· ···················· ····················· ···················· ···················· ····················
P 5885 hCoV-19/England/ALDP-AA1E76/2020|EPI_ISL_625098|20 ···················· ···················· ····················· ···················· ···················· ····················
P 5886 hCoV-19/England/ALDP-AA0E59/2020|EPI_ISL_625099|20 ···················· ···················· ····················· ···················· ···················· ····················
P 5887 hCoV-19/Bahrain/023287015_S14_L001/2020|EPI_ISL_63 ···················· ···················· ····················· ···················· ···················· ····················
P 5888 hCoV-19/Bahrain/090000572_S16_L001/2020|EPI_ISL_63 ···················· ···················· ····················· ···················· ···················· ····················
P 5889 hCoV-19/Faroe Islands/HFS-140/2020|EPI_ISL_614304| ···················· ···················· ····················· ···················· ···················· ····················
P 5890 hCoV-19/Faroe Islands/HFS-51/2020|EPI_ISL_614307|2 ···················· ···················· ····················· ···················· ···················· ····················
P 5891 hCoV-19/Faroe Islands/HFS-60/2020|EPI_ISL_614308|2 ···················· ···················· ····················· ···················· ···················· ····················
P 5892 hCoV-19/Faroe Islands/HFS-71/2020|EPI_ISL_614310|2 ···················· ···················· ····················· ···················· ···················· ····················
P 5893 hCoV-19/Faroe Islands/HFS-74/2020|EPI_ISL_614311|2 ···················· ···················· ····················· ···················· ···················· ····················
P 5894 hCoV-19/Faroe Islands/HFS-77/2020|EPI_ISL_614312|2 ···················· ···················· ····················· ···················· ···················· ····················
P 5895 hCoV-19/Cote dIvoire/C01-2459/2020|EPI_ISL_614347| ···················· ···················· ····················· ···················· ···················· ····················
P 5896 hCoV-19/Cote dIvoire/BKE0011/2020|EPI_ISL_614348|2 ···················· ···················· ····················· ···················· ···················· ····················
P 5897 hCoV-19/Cote dIvoire/BKE0016/2020|EPI_ISL_614349|2 ···················· ···················· ····················· ···················· ···················· ····················
P 5898 hCoV-19/Cote dIvoire/BKE0041/2020|EPI_ISL_614351|2 ···················· ···················· ····················· ···················· ···················· ····················
P 5899 hCoV-19/Cote dIvoire/BKE0042/2020|EPI_ISL_614352|2 ···················· ···················· ····················· ···················· ···················· ····················
P 5900 hCoV-19/Cote dIvoire/BKE0104/2020|EPI_ISL_614353|2 ···················· ···················· ····················· ···················· ···················· ····················
P 5901 hCoV-19/Cote dIvoire/BKE0117/2020|EPI_ISL_614354|2 ···················· ···················· ····················· ···················· ···················· ····················
P 5902 hCoV-19/Cote dIvoire/BKE0125/2020|EPI_ISL_614355|2 ···················· ···················· ····················· ···················· ···················· ····················
P 5903 hCoV-19/Cote dIvoire/BKE0165/2020|EPI_ISL_614356|2 ···················· ···················· ····················· ···················· ···················· ····················
P 5904 hCoV-19/Cote dIvoire/BKE0172/2020|EPI_ISL_614357|2 ···················· ···················· ····················· ···················· ···················· ····················
P 5905 hCoV-19/Cote dIvoire/BKE0173/2020|EPI_ISL_614358|2 ···················· ···················· ····················· ···················· ···················· ····················
P 5906 hCoV-19/Cote dIvoire/BKE0178/2020|EPI_ISL_614359|2 ···················· ···················· ····················· ···················· ···················· ····················
P 5907 hCoV-19/Denmark/DCGC-3889/2020|EPI_ISL_615608|2020 ···················· ···················· ····················· ···················· ···················· ····················
P 5908 hCoV-19/Denmark/DCGC-3890/2020|EPI_ISL_615609|2020 ···················· ···················· ····················· ···················· ···················· ····················
P 5909 hCoV-19/Denmark/DCGC-3891/2020|EPI_ISL_615610|2020 ···················· ···················· ····················· ···················· ···················· ····················
P 5910 hCoV-19/Denmark/DCGC-3892/2020|EPI_ISL_615611|2020 ···················· ···················· ····················· ···················· ···················· ····················
P 5911 hCoV-19/Denmark/DCGC-3894/2020|EPI_ISL_615613|2020 ···················· ···················· ····················· ···················· ···················· ····················
P 5912 hCoV-19/Denmark/DCGC-3895/2020|EPI_ISL_615614|2020 ···················· ···················· ····················· ···················· ···················· ····················
P 5913 hCoV-19/Denmark/DCGC-3896/2020|EPI_ISL_615615|2020 ···················· ···················· ····················· ···················· ···················· ····················
P 5914 hCoV-19/Denmark/DCGC-3897/2020|EPI_ISL_615616|2020 ···················· ···A················ ····················· ···················· ···················· ····················
P 5915 hCoV-19/Denmark/DCGC-1719/2020|EPI_ISL_615617|2020 ···················· ···················· ····················· ···················· ···················· ····················
P 5916 hCoV-19/Denmark/DCGC-3898/2020|EPI_ISL_615618|2020 ···················· ···················· ····················· ···················· ···················· ····················
P 5917 hCoV-19/Denmark/DCGC-3900/2020|EPI_ISL_615620|2020 ···················· ···················· ····················· ···················· ···················· ····················
P 5918 hCoV-19/Denmark/DCGC-3901/2020|EPI_ISL_615621|2020 ··················T· ···················· ····················· ···················· ···················· ···················· 1
P 5919 hCoV-19/Denmark/DCGC-3902/2020|EPI_ISL_615622|2020 ···················· ···················· ····················· ···················· ···················· ····················
P 5920 hCoV-19/Denmark/DCGC-3903/2020|EPI_ISL_615623|2020 ···················· ···················· ····················· ···················· ···················· ····················
P 5921 hCoV-19/Denmark/DCGC-6318/2020|EPI_ISL_615624|2020 ···················· ···················· ····················· ···················· ···················· ····················
P 5922 hCoV-19/Denmark/DCGC-3904/2020|EPI_ISL_615625|2020 ···················· ···················· ····················· ···················· ···················· ····················
P 5923 hCoV-19/Denmark/DCGC-3905/2020|EPI_ISL_615626|2020 ···················· ···················· ····················· ···················· ···················· ····················
P 5924 hCoV-19/Denmark/DCGC-3906/2020|EPI_ISL_615627|2020 ···················· ···················· ····················· ···················· ···················· ····················
P 5925 hCoV-19/Denmark/DCGC-3907/2020|EPI_ISL_615628|2020 ···················· ···················· ····················· ···················· ···················· ····················
P 5926 hCoV-19/Denmark/DCGC-3908/2020|EPI_ISL_615629|2020 ···················· ···················· ····················· ···················· ···················· ····················
P 5927 hCoV-19/Denmark/DCGC-3909/2020|EPI_ISL_615630|2020 ···················· ···················· ····················· ···················· ···················· ····················
P 5928 hCoV-19/Denmark/DCGC-3910/2020|EPI_ISL_615631|2020 ···················· ···················· ····················· ···················· ···················· ····················
P 5929 hCoV-19/Denmark/DCGC-3911/2020|EPI_ISL_615632|2020 ···················· ···················· ····················· ···················· ···················· ····················
P 5930 hCoV-19/Denmark/DCGC-3912/2020|EPI_ISL_615633|2020 ···················· ···················· ····················· ···················· ···················· ····················
P 5931 hCoV-19/Denmark/DCGC-3913/2020|EPI_ISL_615634|2020 ···················· ···················· ····················· ···················· ···················· ····················
P 5932 hCoV-19/Denmark/DCGC-3914/2020|EPI_ISL_615635|2020 ···················· ···················· ····················· ···················· ···················· ····················
P 5933 hCoV-19/Denmark/DCGC-1716/2020|EPI_ISL_615637|2020 ···················· ···················· ····················· ···················· ···················· ····················
P 5934 hCoV-19/Denmark/DCGC-4453/2020|EPI_ISL_615638|2020 ···················· ···················· ····················· ···················· ···················· ····················
P 5935 hCoV-19/Denmark/DCGC-4454/2020|EPI_ISL_615639|2020 ···················· ···················· ····················· ···················· ···················· ····················
P 5936 hCoV-19/Denmark/DCGC-4455/2020|EPI_ISL_615640|2020 ···················· ···················· ····················· ···················· ···················· ····················
P 5937 hCoV-19/Denmark/DCGC-4456/2020|EPI_ISL_615641|2020 ···················· ···················· ····················· ···················· ···················· ····················
P 5938 hCoV-19/Denmark/DCGC-4457/2020|EPI_ISL_615642|2020 ···················· ···················· ····················· ···················· ···················· ····················
P 5939 hCoV-19/Denmark/DCGC-4458/2020|EPI_ISL_615643|2020 ···················· ·A·················· ····················· ···················· ···················· ····················
P 5940 hCoV-19/Denmark/DCGC-4459/2020|EPI_ISL_615644|2020 ···················· ·A·················· ····················· ···················· ···················· ····················
P 5941 hCoV-19/Denmark/DCGC-4460/2020|EPI_ISL_615645|2020 ···················· ···················· ····················· ···················· ···················· ····················
P 5942 hCoV-19/Denmark/DCGC-4461/2020|EPI_ISL_615646|2020 ···················· ···················· ····················· ···················· ···················· ····················
P 5943 hCoV-19/Denmark/DCGC-4462/2020|EPI_ISL_615647|2020 ···················· ···················· ····················· ···················· ···················· ····················
P 5944 hCoV-19/Denmark/DCGC-4463/2020|EPI_ISL_615648|2020 ···················· ···················· ····················· ···················· ···················· ····················
P 5945 hCoV-19/Denmark/DCGC-4464/2020|EPI_ISL_615649|2020 ···················· ···················· ····················· ···················· ···················· ····················
P 5946 hCoV-19/Denmark/DCGC-4465/2020|EPI_ISL_615650|2020 ···················· ···················· ····················· ···················· ···················· ····················
P 5947 hCoV-19/Denmark/DCGC-4469/2020|EPI_ISL_615654|2020 ···················· ···················· ····················· ···················· ···················· ····················
P 5948 hCoV-19/Denmark/DCGC-4470/2020|EPI_ISL_615655|2020 ···················· ···················· ····················· ···················· ···················· ····················
P 5949 hCoV-19/Denmark/DCGC-4471/2020|EPI_ISL_615656|2020 ···················· ···················· ····················· ···················· ···················· ····················
P 5950 hCoV-19/Denmark/DCGC-4473/2020|EPI_ISL_615658|2020 ···················· ···················· ····················· ···················· ···················· ····················
P 5951 hCoV-19/Denmark/DCGC-4474/2020|EPI_ISL_615659|2020 ···················· ···················· ····················· ···················· ···················· ····················
P 5952 hCoV-19/Denmark/DCGC-4475/2020|EPI_ISL_615660|2020 ···················· ···················· ····················· ···················· ···················· ····················
P 5953 hCoV-19/Denmark/DCGC-4477/2020|EPI_ISL_615662|2020 ···················· ···················· ····················· ···················· ···················· ····················
P 5954 hCoV-19/Denmark/DCGC-4478/2020|EPI_ISL_615663|2020 ···················· ···················· ····················· ···················· ···················· ····················
P 5955 hCoV-19/Denmark/DCGC-4479/2020|EPI_ISL_615664|2020 ···················· ···················· ····················· ···················· ···················· ····················
P 5956 hCoV-19/Denmark/DCGC-4480/2020|EPI_ISL_615665|2020 ···················· ···················· ····················· ···················· ···················· ····················
P 5957 hCoV-19/Denmark/DCGC-4481/2020|EPI_ISL_615666|2020 ···················· ···················· ····················· ···················· ···················· ····················
P 5958 hCoV-19/Denmark/DCGC-4483/2020|EPI_ISL_615668|2020 ···················· ···················· ····················· ···················· ···················· ····················
P 5959 hCoV-19/Denmark/DCGC-4484/2020|EPI_ISL_615669|2020 ···················· ···················· ····················· ···················· ···················· ····················
P 5960 hCoV-19/Denmark/DCGC-4485/2020|EPI_ISL_615670|2020 ···················· ···················· ····················· ···················· ···················· ····················
```

| | | | | | | | | |
|---|---|---|---|---|---|---|---|---|
| P | 5961 | hCoV-19/Denmark/DCGC-4486/2020\|EPI_ISL_615671\|2020 | ···················· | ···················· | ····················· | ···················· | ···················· | ···················· |
| P | 5962 | hCoV-19/Denmark/DCGC-4487/2020\|EPI_ISL_615672\|2020 | ···················· | ···················· | ····················· | ···················· | ···················· | ···················· |
| P | 5963 | hCoV-19/Denmark/DCGC-4489/2020\|EPI_ISL_615674\|2020 | ···················· | ···················· | ····················· | ···················· | ···················· | ···················· |
| P | 5964 | hCoV-19/Denmark/DCGC-4490/2020\|EPI_ISL_615675\|2020 | ···················· | ···················· | ····················· | ···················· | ···················· | ···················· |
| P | 5965 | hCoV-19/Denmark/DCGC-4491/2020\|EPI_ISL_615676\|2020 | ···················· | ···················· | ····················· | ···················· | ···················· | ···················· |
| P | 5966 | hCoV-19/Denmark/DCGC-4492/2020\|EPI_ISL_615677\|2020 | ···················· | ···················· | ····················· | ···················· | ···················· | ···················· |
| P | 5967 | hCoV-19/Denmark/DCGC-4494/2020\|EPI_ISL_615679\|2020 | ···················· | ···················· | ····················· | ···················· | ···················· | ···················· |
| P | 5968 | hCoV-19/Denmark/DCGC-4495/2020\|EPI_ISL_615680\|2020 | ···················· | ···················· | ····················· | ···················· | ···················· | ···················· |
| P | 5969 | hCoV-19/Denmark/DCGC-4496/2020\|EPI_ISL_615681\|2020 | ···················· | ···················· | ····················· | ···················· | ···················· | ···················· |
| P | 5970 | hCoV-19/Denmark/DCGC-4497/2020\|EPI_ISL_615682\|2020 | ···················· | ···················· | ····················· | ···················· | ···················· | ···················· |
| P | 5971 | hCoV-19/Denmark/DCGC-4498/2020\|EPI_ISL_615683\|2020 | ···················· | ···················· | ····················· | ···················· | ···················· | ···················· |
| P | 5972 | hCoV-19/Denmark/DCGC-4500/2020\|EPI_ISL_615685\|2020 | ···················· | ···················· | ····················· | ···················· | ···················· | ···················· |
| P | 5973 | hCoV-19/Denmark/DCGC-4501/2020\|EPI_ISL_615686\|2020 | ···················· | ···················· | ····················· | ···················· | ···················· | ···················· |
| P | 5974 | hCoV-19/Denmark/DCGC-4502/2020\|EPI_ISL_615687\|2020 | ···················· | ···················· | ····················· | ···················· | ···················· | ···················· |
| P | 5975 | hCoV-19/Denmark/DCGC-4504/2020\|EPI_ISL_615689\|2020 | ···················· | ···················· | ····················· | ···················· | ···················· | ···················· |
| P | 5976 | hCoV-19/Denmark/DCGC-8469/2020\|EPI_ISL_616951\|2020 | ···················· | ···················· | ····················· | ···················· | ···················· | ···················· |
| P | 5977 | hCoV-19/Denmark/DCGC-8002/2020\|EPI_ISL_616952\|2020 | ···················· | ···················· | ····················· | ···················· | ···················· | ···················· |
| P | 5978 | hCoV-19/Denmark/DCGC-8018/2020\|EPI_ISL_616953\|2020 | ···················· | ···················· | ····················· | ···················· | ···················· | ···················· |
| P | 5979 | hCoV-19/Denmark/DCGC-7963/2020\|EPI_ISL_616954\|2020 | ···················· | ···················· | ····················· | ···················· | ···················· | ···················· |
| P | 5980 | hCoV-19/Denmark/DCGC-8136/2020\|EPI_ISL_616955\|2020 | ···················· | ···················· | ····················· | ···················· | ···················· | ···················· |
| P | 5981 | hCoV-19/Denmark/DCGC-8256/2020\|EPI_ISL_616956\|2020 | ···················· | ···················· | ····················· | ···················· | ···················· | ···················· |
| P | 5982 | hCoV-19/Denmark/DCGC-8273/2020\|EPI_ISL_616976\|2020 | ···················· | ······A············· | ····················· | ···················· | ···················· | ···················· |
| P | 5983 | hCoV-19/Denmark/DCGC-7935/2020\|EPI_ISL_616977\|2020 | ···················· | ···················· | ····················· | ···················· | ···················· | ···················· |
| P | 5984 | hCoV-19/Denmark/DCGC-7964/2020\|EPI_ISL_616978\|2020 | ···················· | ···················· | ····················· | ···················· | ···················· | ···················· |
| P | 5985 | hCoV-19/Denmark/DCGC-7924/2020\|EPI_ISL_616979\|2020 | ···················· | ···················· | ····················· | ···················· | ···················· | ···················· |
| P | 5986 | hCoV-19/Denmark/DCGC-7965/2020\|EPI_ISL_616980\|2020 | ···················· | ···················· | ····················· | ···················· | ···················· | ···················· |
| P | 5987 | hCoV-19/Denmark/DCGC-7976/2020\|EPI_ISL_616981\|2020 | ···················· | ···················· | ····················· | ···················· | ···················· | ···················· |
| P | 5988 | hCoV-19/Denmark/DCGC-8037/2020\|EPI_ISL_616982\|2020 | ···················· | ···················· | ····················· | ···················· | ···················· | ···················· |
| P | 5989 | hCoV-19/Denmark/DCGC-8038/2020\|EPI_ISL_616983\|2020 | ···················· | ···················· | ····················· | ···················· | ···················· | ···················· |
| P | 5990 | hCoV-19/Denmark/DCGC-8003/2020\|EPI_ISL_616984\|2020 | ···················· | ···················· | ····················· | ···················· | ···················· | ···················· |
| P | 5991 | hCoV-19/Denmark/DCGC-8040/2020\|EPI_ISL_616985\|2020 | ···················· | ···················· | ····················· | ···················· | ···················· | ···················· |
| P | 5992 | hCoV-19/Denmark/DCGC-7979/2020\|EPI_ISL_616986\|2020 | ···················· | ···················· | ····················· | ···················· | ···················· | ···················· |
| P | 5993 | hCoV-19/Denmark/DCGC-8026/2020\|EPI_ISL_616987\|2020 | ···················· | ···················· | ····················· | ···················· | ···················· | ···················· |
| P | 5994 | hCoV-19/Denmark/DCGC-8041/2020\|EPI_ISL_616988\|2020 | ···················· | ···················· | ····················· | ···················· | ···················· | ···················· |
| P | 5995 | hCoV-19/Denmark/DCGC-8017/2020\|EPI_ISL_616989\|2020 | ···················· | ···················· | ····················· | ···················· | ···················· | ···················· |
| P | 5996 | hCoV-19/Denmark/DCGC-7998/2020\|EPI_ISL_616990\|2020 | ···················· | ···················· | ····················· | ···················· | ···················· | ···················· |
| P | 5997 | hCoV-19/Denmark/DCGC-7999/2020\|EPI_ISL_616991\|2020 | ···················· | ···················· | ····················· | ···················· | ···················· | ···················· |
| P | 5998 | hCoV-19/Denmark/DCGC-8088/2020\|EPI_ISL_616992\|2020 | ···················· | ···················· | ····················· | ···················· | ···················· | ···················· |
| P | 5999 | hCoV-19/Denmark/DCGC-8089/2020\|EPI_ISL_616993\|2020 | ···················· | ···················· | ····················· | ···················· | ···················· | ···················· |
| P | 6000 | hCoV-19/Denmark/DCGC-8107/2020\|EPI_ISL_616994\|2020 | ···················· | ···················· | ····················· | ···················· | ···················· | ···················· |
| P | 6001 | hCoV-19/Denmark/DCGC-8044/2020\|EPI_ISL_616995\|2020 | ···················· | ···················· | ····················· | ···················· | ···················· | ···················· |
| P | 6002 | hCoV-19/Denmark/DCGC-8046/2020\|EPI_ISL_616996\|2020 | ···················· | ···················· | ····················· | ···················· | ···················· | ···················· |
| P | 6003 | hCoV-19/Denmark/DCGC-8104/2020\|EPI_ISL_616997\|2020 | ···················· | ···················· | ····················· | ···················· | ···················· | ···················· |
| P | 6004 | hCoV-19/Denmark/DCGC-8080/2020\|EPI_ISL_616998\|2020 | ···················· | ···················· | ····················· | ···················· | ···················· | ···················· |
| P | 6005 | hCoV-19/Denmark/DCGC-8067/2020\|EPI_ISL_616999\|2020 | ···················· | ···················· | ····················· | ···················· | ···················· | ···················· |
| P | 6006 | hCoV-19/Denmark/DCGC-1629/2020\|EPI_ISL_619260\|2020 | ···················· | ···················· | ····················· | ···················· | ···················· | ···················· |
| P | 6007 | hCoV-19/Denmark/DCGC-1628/2020\|EPI_ISL_619261\|2020 | ···················· | ·A·················· | ····················· | ···················· | ···················· | ···················· |
| P | 6008 | hCoV-19/Denmark/DCGC-1627/2020\|EPI_ISL_619262\|2020 | ···················· | ···················· | ····················· | ···················· | ···················· | ···················· |
| P | 6009 | hCoV-19/Denmark/DCGC-1626/2020\|EPI_ISL_619263\|2020 | ···················· | ···················· | ····················· | ···················· | ···················· | ···················· |
| P | 6010 | hCoV-19/Denmark/DCGC-2051/2020\|EPI_ISL_619264\|2020 | ···················· | ···················· | ····················· | ···················· | ···················· | ···················· |
| P | 6011 | hCoV-19/Denmark/DCGC-1625/2020\|EPI_ISL_619265\|2020 | ···················· | ···················· | ····················· | ···················· | ···················· | ···················· |
| P | 6012 | hCoV-19/Denmark/DCGC-1624/2020\|EPI_ISL_619266\|2020 | ···················· | ···················· | ····················· | ···················· | ···················· | ···················· |
| P | 6013 | hCoV-19/Denmark/DCGC-1623/2020\|EPI_ISL_619267\|2020 | ···················· | ···················· | ····················· | ···················· | ···················· | ···················· |
| P | 6014 | hCoV-19/Denmark/DCGC-1622/2020\|EPI_ISL_619268\|2020 | ···················· | ···················· | ····················· | ···················· | ···················· | ···················· |
| P | 6015 | hCoV-19/Denmark/DCGC-1620/2020\|EPI_ISL_619269\|2020 | ···················· | ···················· | ····················· | ···················· | ···················· | ···················· |
| P | 6016 | hCoV-19/Denmark/DCGC-2053/2020\|EPI_ISL_619270\|2020 | ···················· | ···················· | ····················· | ···················· | ···················· | ···················· |
| P | 6017 | hCoV-19/Denmark/DCGC-1619/2020\|EPI_ISL_619271\|2020 | ···················· | ···················· | ····················· | ···················· | ···················· | ···················· |
| P | 6018 | hCoV-19/Denmark/DCGC-1618/2020\|EPI_ISL_619272\|2020 | ···················· | ···················· | ····················· | ···················· | ···················· | ···················· |
| P | 6019 | hCoV-19/Denmark/DCGC-1617/2020\|EPI_ISL_619273\|2020 | ···················· | ···················· | ····················· | ···················· | ···················· | ···················· |
| P | 6020 | hCoV-19/Denmark/DCGC-7306/2020\|EPI_ISL_619274\|2020 | ···················· | ···················· | ····················· | ···················· | ···················· | ···················· |
| P | 6021 | hCoV-19/Denmark/DCGC-7332/2020\|EPI_ISL_619275\|2020 | ···················· | ···················· | ····················· | ···················· | ···················· | ···················· |
| P | 6022 | hCoV-19/Denmark/DCGC-7340/2020\|EPI_ISL_619276\|2020 | ···················· | ···················· | ····················· | ···················· | ···················· | ···················· |
| P | 6023 | hCoV-19/Denmark/DCGC-7368/2020\|EPI_ISL_619277\|2020 | ···················· | ···················· | ····················· | ···················· | ···················· | ···················· |
| P | 6024 | hCoV-19/Denmark/DCGC-7375/2020\|EPI_ISL_619278\|2020 | ···················· | ···················· | ····················· | ···················· | ···················· | ···················· |
| P | 6025 | hCoV-19/Denmark/DCGC-7307/2020\|EPI_ISL_619279\|2020 | ···················· | ···················· | ····················· | ···················· | ···················· | ···················· |
| P | 6026 | hCoV-19/Denmark/DCGC-7315/2020\|EPI_ISL_619280\|2020 | ···················· | ···················· | ····················· | ···················· | ···················· | ···················· |
| P | 6027 | hCoV-19/Denmark/DCGC-7327/2020\|EPI_ISL_619281\|2020 | ···················· | ···················· | ····················· | ···················· | ···················· | ···················· |
| P | 6028 | hCoV-19/Denmark/DCGC-7333/2020\|EPI_ISL_619282\|2020 | ···················· | ···················· | ····················· | ···················· | ···················· | ···················· |
| P | 6029 | hCoV-19/Denmark/DCGC-7341/2020\|EPI_ISL_619283\|2020 | ···················· | ···················· | ····················· | ···················· | ···················· | ···················· |
| P | 6030 | hCoV-19/Denmark/DCGC-7349/2020\|EPI_ISL_619284\|2020 | ···················· | ···················· | ····················· | ···················· | ···················· | ···················· |
| P | 6031 | hCoV-19/Denmark/DCGC-7369/2020\|EPI_ISL_619285\|2020 | ···················· | ···················· | ····················· | ···················· | ···················· | ···················· |
| P | 6032 | hCoV-19/Denmark/DCGC-7376/2020\|EPI_ISL_619286\|2020 | ···················· | ···················· | ····················· | ···················· | ···················· | ···················· |
| P | 6033 | hCoV-19/Denmark/DCGC-7308/2020\|EPI_ISL_619287\|2020 | ···················· | ···················· | ····················· | ···················· | ···················· | ···················· |
| P | 6034 | hCoV-19/Denmark/DCGC-7328/2020\|EPI_ISL_619288\|2020 | ···················· | ····A··············· | ····················· | ···················· | ···················· | ···················· |
| P | 6035 | hCoV-19/Denmark/DCGC-7342/2020\|EPI_ISL_619289\|2020 | ···················· | ···················· | ····················· | ···················· | ···················· | ···················· |
| P | 6036 | hCoV-19/Denmark/DCGC-6043/2020\|EPI_ISL_620237\|2020 | ···················· | ···················· | ····················· | ···················· | ···················· | ···················· |
| P | 6037 | hCoV-19/Denmark/DCGC-6044/2020\|EPI_ISL_620238\|2020 | ···················· | ···················· | ····················· | ···················· | ···················· | ···················· |
| P | 6038 | hCoV-19/Denmark/DCGC-6045/2020\|EPI_ISL_620239\|2020 | ···················· | ···················· | ····················· | ···················· | ···················· | ···················· |
| P | 6039 | hCoV-19/Denmark/DCGC-6046/2020\|EPI_ISL_620240\|2020 | ···················· | ···················· | ····················· | ···················· | ···················· | ···················· |
| P | 6040 | hCoV-19/Denmark/DCGC-6047/2020\|EPI_ISL_620241\|2020 | ···················· | ···················· | ····················· | ···················· | ···················· | ···················· |

P 6041 hCoV-19/Denmark/DCGC-6048/2020|EPI_ISL_620242|2020 ···················· ···················· ····················· ···················· ···················· ····················
P 6042 hCoV-19/Denmark/DCGC-6049/2020|EPI_ISL_620243|2020 ···················· ···················· ····················· ···················· ···················· ····················
P 6043 hCoV-19/Denmark/DCGC-6050/2020|EPI_ISL_620244|2020 ···················· ···················· ····················· ···················· ···················· ····················
P 6044 hCoV-19/Denmark/DCGC-6051/2020|EPI_ISL_620245|2020 ···················· ···················· ····················· ···················· ···················· ····················
P 6045 hCoV-19/Denmark/DCGC-6052/2020|EPI_ISL_620246|2020 ···················· ···················· ····················· ···················· ···················· ····················
P 6046 hCoV-19/Denmark/DCGC-6053/2020|EPI_ISL_620247|2020 ···················· ···················· ····················· ···················· ···················· ····················
P 6047 hCoV-19/Denmark/DCGC-6057/2020|EPI_ISL_620251|2020 ···················· ···················· ····················· ···················· ···················· ····················
P 6048 hCoV-19/Denmark/DCGC-6058/2020|EPI_ISL_620252|2020 ···················· ···················· ····················· ···················· ···················· ····················
P 6049 hCoV-19/Denmark/DCGC-6060/2020|EPI_ISL_620254|2020 ···················· ···················· ····················· ···················· ···················· ····················
P 6050 hCoV-19/Denmark/DCGC-6061/2020|EPI_ISL_620255|2020 ···················· ···················· ····················· ···················· ···················· ····················
P 6051 hCoV-19/Denmark/DCGC-6062/2020|EPI_ISL_620256|2020 ···················· ···················· ····················· ···················· ···················· ····················
P 6052 hCoV-19/Denmark/DCGC-6063/2020|EPI_ISL_620257|2020 ···················· ···················· ····················· ···················· ···················· ····················
P 6053 hCoV-19/Denmark/DCGC-6065/2020|EPI_ISL_620259|2020 ···················· ···················· ····················· ···················· ···················· ····················
P 6054 hCoV-19/Denmark/DCGC-6066/2020|EPI_ISL_620260|2020 ···················· ···················· ····················· ···················· ···················· ····················
P 6055 hCoV-19/Denmark/DCGC-6067/2020|EPI_ISL_620261|2020 ···················· ···················· ····················· ···················· ···················· ····················
P 6056 hCoV-19/Denmark/DCGC-6068/2020|EPI_ISL_620262|2020 ···················· ···················· ····················· ···················· ···················· ····················
P 6057 hCoV-19/Denmark/DCGC-6069/2020|EPI_ISL_620263|2020 ···················· ···················· ····················· ···················· ···················· ····················
P 6058 hCoV-19/Denmark/DCGC-6070/2020|EPI_ISL_620264|2020 ···················· ·A·················· ····················· ···················· ···················· ····················
P 6059 hCoV-19/Denmark/DCGC-6072/2020|EPI_ISL_620266|2020 ···················· ···················· ····················· ···················· ···················· ····················
P 6060 hCoV-19/Denmark/DCGC-6074/2020|EPI_ISL_620268|2020 ···················· ···················· ····················· ···················· ···················· ····················
P 6061 hCoV-19/Denmark/DCGC-6075/2020|EPI_ISL_620269|2020 ···················· ···················· ····················· ···················· ···················· ····················
P 6062 hCoV-19/Denmark/DCGC-6076/2020|EPI_ISL_620270|2020 ···················· ···················· ····················· ···················· ···················· ····················
P 6063 hCoV-19/Denmark/DCGC-6077/2020|EPI_ISL_620271|2020 ···················· ···················· ····················· ···················· ···················· ····················
P 6064 hCoV-19/Denmark/DCGC-6078/2020|EPI_ISL_620272|2020 ···················· ···················· ····················· ···················· ···················· ····················
P 6065 hCoV-19/Denmark/DCGC-6079/2020|EPI_ISL_620273|2020 ···················· ···················· ····················· ···················· ···················· ····················
P 6066 hCoV-19/Denmark/DCGC-6080/2020|EPI_ISL_620274|2020 ···················· ···················· ····················· ···················· ···················· ····················
P 6067 hCoV-19/Denmark/DCGC-6081/2020|EPI_ISL_620275|2020 ···················· ···················· ····················· ···················· ···················· ····················
P 6068 hCoV-19/Denmark/DCGC-6083/2020|EPI_ISL_620277|2020 ···················· ···················· ····················· ···················· ···················· ····················
P 6069 hCoV-19/Denmark/DCGC-6084/2020|EPI_ISL_620278|2020 ···················· ···················· ····················· ···················· ···················· ····················
P 6070 hCoV-19/Denmark/DCGC-6085/2020|EPI_ISL_620279|2020 ···················· ···················· ····················· ···················· ···················· ····················
P 6071 hCoV-19/Denmark/DCGC-4882/2020|EPI_ISL_621568|2020 ···················· ···················· ····················· ···················· ···················· ····················
P 6072 hCoV-19/Denmark/DCGC-4883/2020|EPI_ISL_621569|2020 ···················· ···················· ····················· ···················· ···················· ····················
P 6073 hCoV-19/Denmark/DCGC-4884/2020|EPI_ISL_621570|2020 ···················· ···················· ····················· ···················· ···················· ····················
P 6074 hCoV-19/Denmark/DCGC-4885/2020|EPI_ISL_621571|2020 ···················· ···················· ····················· ···················· ···················· ····················
P 6075 hCoV-19/Denmark/DCGC-4886/2020|EPI_ISL_621572|2020 ···················· ···················· ····················· ···················· ···················· ····················
P 6076 hCoV-19/Denmark/DCGC-4887/2020|EPI_ISL_621573|2020 ···················· ···················· ····················· ···················· ···················· ····················
P 6077 hCoV-19/Denmark/DCGC-4888/2020|EPI_ISL_621574|2020 ···················· ···················· ····················· ···················· ···················· ····················
P 6078 hCoV-19/Denmark/DCGC-4889/2020|EPI_ISL_621575|2020 ···················· ···················· ····················· ···················· ···················· ····················
P 6079 hCoV-19/Denmark/DCGC-4890/2020|EPI_ISL_621576|2020 ···················· ···················· ····················· ···················· ···················· ····················
P 6080 hCoV-19/Denmark/DCGC-4891/2020|EPI_ISL_621577|2020 ···················· ···················· ····················· ···················· ···················· ····················
P 6081 hCoV-19/Denmark/DCGC-4892/2020|EPI_ISL_621578|2020 ···················· ···················· ····················· ···················· ···················· ····················
P 6082 hCoV-19/Denmark/DCGC-4896/2020|EPI_ISL_621582|2020 ···················· ···················· ····················· ···················· ···················· ····················
P 6083 hCoV-19/Denmark/DCGC-4897/2020|EPI_ISL_621583|2020 ···················· ···················· ····················· ···················· ···················· ····················
P 6084 hCoV-19/Denmark/DCGC-4898/2020|EPI_ISL_621584|2020 ···················· ···················· ····················· ···················· ···················· ····················
P 6085 hCoV-19/Denmark/DCGC-4899/2020|EPI_ISL_621585|2020 ···················· ·A·················· ····················· ···················· ···················· ····················
P 6086 hCoV-19/Denmark/DCGC-4900/2020|EPI_ISL_621586|2020 ···················· ···················· ····················· ···················· ···················· ····················
P 6087 hCoV-19/Denmark/DCGC-4901/2020|EPI_ISL_621587|2020 ···················· ···················· ····················· ···················· ···················· ····················
P 6088 hCoV-19/Denmark/DCGC-5052/2020|EPI_ISL_621590|2020 ···················· ···················· ····················· ···················· ···················· ····················
P 6089 hCoV-19/Denmark/DCGC-5053/2020|EPI_ISL_621591|2020 ···················· ···················· ····················· ···················· ···················· ····················
P 6090 hCoV-19/Denmark/DCGC-5056/2020|EPI_ISL_621594|2020 ···················· ···················· ····················· ···················· ···················· ····················
P 6091 hCoV-19/Denmark/DCGC-5057/2020|EPI_ISL_621595|2020 ···················· ···················· ····················· ···················· ···················· ····················
P 6092 hCoV-19/Denmark/DCGC-5059/2020|EPI_ISL_621597|2020 ···················· ···················· ····················· ···················· ···················· ····················
P 6093 hCoV-19/Denmark/DCGC-5060/2020|EPI_ISL_621598|2020 ···················· ···················· ····················· ···················· ···················· ····················
P 6094 hCoV-19/Denmark/DCGC-5061/2020|EPI_ISL_621599|2020 ···················· ···················· ····················· ···················· ···················· ····················
P 6095 hCoV-19/Wales/ALDP-AA0949/2020|EPI_ISL_625000|2020 ···················· ···················· ····················· ···················· ···················· ····················
P 6096 hCoV-19/England/ALDP-AA218B/2020|EPI_ISL_625001|20 ···················· ···················· ····················· ···················· ···················· ····················
P 6097 hCoV-19/England/ALDP-AA37F1/2020|EPI_ISL_625002|20 ···················· ···················· ····················· ···················· ···················· ····················
P 6098 hCoV-19/England/ALDP-AA17C6/2020|EPI_ISL_625004|20 ···················· ···················· ····················· ···················· ···················· ····················
P 6099 hCoV-19/England/ALDP-AA365E/2020|EPI_ISL_625005|20 ···················· ···················· ····················· ···················· ···················· ····················
P 6100 hCoV-19/England/ALDP-AA093A/2020|EPI_ISL_625007|20 ···················· ···················· ····················· ···················· ···················· ····················
P 6101 hCoV-19/England/ALDP-AA115F/2020|EPI_ISL_625008|20 ···················· ···················· ····················· ···················· ···················· ····················
P 6102 hCoV-19/England/ALDP-AA32D2/2020|EPI_ISL_625009|20 ···················· ···················· ····················· ···················· ···················· ····················
P 6103 hCoV-19/England/ALDP-AA5373/2020|EPI_ISL_625060|20 ···················· ···················· ····················· ···················· ···················· ····················
P 6104 hCoV-19/England/ALDP-AA1E3A/2020|EPI_ISL_625061|20 ···················· ·A·················· ····················· ···················· ···················· ··A·················
P 6105 hCoV-19/England/ALDP-AA37C4/2020|EPI_ISL_625062|20 ···················· ···················· ····················· ···················· ···················· ····················
P 6106 hCoV-19/England/ALDP-AA11B9/2020|EPI_ISL_625063|20 ···················· ···················· ····················· ···················· ···················· ····················
P 6107 hCoV-19/Wales/ALDP-AA37D3/2020|EPI_ISL_625064|2020 ···················· ···················· ····················· ···················· ···················· ····················
P 6108 hCoV-19/Wales/ALDP-AA49AF/2020|EPI_ISL_625065|2020 ···················· ···················· ····················· ···················· ···················· ····················
P 6109 hCoV-19/England/ALDP-AA20E8/2020|EPI_ISL_625069|20 ···················· ···················· ····················· ···················· ···················· ····················
P 6110 hCoV-19/Belgium/ULG-10529/2020|EPI_ISL_626300|2020 ···················· ···················· ····················· ···················· ···················· ····················
P 6111 hCoV-19/Belgium/ULG-10526/2020|EPI_ISL_626301|2020 ···················· ···················· ····················· ···················· ···················· ····················
P 6112 hCoV-19/Belgium/ULG-10587/2020|EPI_ISL_626302|2020 ···················· ···················· ····················· ···················· ···················· ····················
P 6113 hCoV-19/Belgium/ULG-10544/2020|EPI_ISL_626303|2020 ···················· ···················· ····················· ···················· ···················· ····················
P 6114 hCoV-19/Belgium/ULG-10552/2020|EPI_ISL_626304|2020 ···················· ···················· ····················· ···················· ···················· ····················
P 6116 hCoV-19/Belgium/ULG-10545/2020|EPI_ISL_626306|2020 ···················· ···················· ····················· ···················· ···················· ····················
P 6117 hCoV-19/Belgium/ULG-10512/2020|EPI_ISL_626307|2020 ···················· ···················· ····················· ···················· ···················· ····················
P 6119 hCoV-19/Belgium/ULG-10592/2020|EPI_ISL_626311|2020 ···················· ···················· ····················· ···················· ···················· ····················
P 6120 hCoV-19/Belgium/ULG-10551/2020|EPI_ISL_626312|2020 ···················· ···················· ····················· ···················· ···················· ····················
P 6121 hCoV-19/Belgium/ULG-10535/2020|EPI_ISL_626314|2020 ···················· ···················· ····················· ···················· ···················· ····················
P 6122 hCoV-19/Belgium/ULG-10531/2020|EPI_ISL_626315|2020 ···················· ···················· ····················· ···················· ···················· ····················

P 6123 hCoV-19/Belgium/ULG-10570/2020|EPI_ISL_626316|2020 .................... .................... .................... .................... .................... ....................
P 6124 hCoV-19/Belgium/ULG-10543/2020|EPI_ISL_626318|2020 .................... .................... .................... .................... .................... ....................
P 6126 hCoV-19/Belgium/ULG-10532/2020|EPI_ISL_626320|2020 .................... .................... .................... .................... .................... ....................
P 6127 hCoV-19/Belgium/ULG-10527/2020|EPI_ISL_626321|2020 .................... .................... .................... .................... .................... ....................
P 6128 hCoV-19/Belgium/ULG-10530/2020|EPI_ISL_626322|2020 .................... .................... .................... .................... .................... ....................
P 6131 hCoV-19/Belgium/ULG-10533/2020|EPI_ISL_626325|2020 .................... .................... .................... .................... .................... ....................
P 6132 hCoV-19/Belgium/ULG-10538/2020|EPI_ISL_626326|2020 .................... .................... .................... .................... .................... ....................
P 6133 hCoV-19/Belgium/ULG-10525/2020|EPI_ISL_626327|2020 .................... .................... .................... .................... .................... ....................
P 6134 hCoV-19/Belgium/ULG-10560/2020|EPI_ISL_626328|2020 .................... .................... .................... .................... .................... ....................
P 6136 hCoV-19/Belgium/ULG-10515/2020|EPI_ISL_626331|2020 .................... .................... .................... .................... .................... ....................
P 6139 hCoV-19/Belgium/ULG-10567/2020|EPI_ISL_626334|2020 .................... .................... .................... .................... .................... ....................
P 6141 hCoV-19/Belgium/ULG-10585/2020|EPI_ISL_626336|2020 .................... .................... .................... .................... .................... ....................
P 6143 hCoV-19/Belgium/ULG-10588/2020|EPI_ISL_626338|2020 .................... .................... .................... .................... .................... ....................
P 6145 hCoV-19/Cote dIvoire/BKE0189/2020|EPI_ISL_614360|2 .................... .................... .................... .................... .................... ....................
P 6147 hCoV-19/Cote dIvoire/BKE0202/2020|EPI_ISL_614362|2 .................... .................... .................... .................... .................... ....................
P 6148 hCoV-19/Cote dIvoire/BKE0213/2020|EPI_ISL_614363|2 .................... .................... .................... .................... .................... ....................
P 6149 hCoV-19/Cote dIvoire/BKE0247/2020|EPI_ISL_614364|2 .................... .................... .................... .................... .................... ....................
P 6150 hCoV-19/Cote dIvoire/BKE0248/2020|EPI_ISL_614365|2 .................... .................... .................... .................... .................... ....................
P 6151 hCoV-19/Cote dIvoire/BKE0249/2020|EPI_ISL_614366|2 .................... .................... .................... .................... .................... ....................
P 6153 hCoV-19/Cote dIvoire/BKE0257/2020|EPI_ISL_614368|2 .................... .................... .................... .................... .................... ....................
P 6154 hCoV-19/Cote dIvoire/BKE0310/2020|EPI_ISL_614371|2 .................... .................... .................... .................... .................... ....................
P 6157 hCoV-19/Cote dIvoire/BKE0424/2020|EPI_ISL_614376|2 .................... .................... .................... .................... .................... ....................
P 6158 hCoV-19/Cote dIvoire/BKE0622/2020|EPI_ISL_614377|2 .................... .................... .................... .................... .................... ....................
P 6159 hCoV-19/Cote dIvoire/BKE0631/2020|EPI_ISL_614379|2 .................... .................... .................... .................... .................... ....................
P 6162 hCoV-19/Cote dIvoire/BKE0753/2020|EPI_ISL_614384|2 .................... .................... .................... .................... .................... ....................
P 6163 hCoV-19/Cote dIvoire/BKE0862/2020|EPI_ISL_614386|2 .................... .................... .................... .................... .................... ....................
P 6165 hCoV-19/Cote dIvoire/BKE0888/2020|EPI_ISL_614388|2 .................... .................... .................... .................... .................... ....................
P 6166 hCoV-19/Cote dIvoire/BKE0916/2020|EPI_ISL_614391|2 .................... .................... .................... .................... .................... ....................
P 6167 hCoV-19/Cote dIvoire/BKE0965/2020|EPI_ISL_614393|2 .................... .................... .................... .................... .................... ....................
P 6168 hCoV-19/Italy/SAR-ASSL-07/2020|EPI_ISL_614396|2020 .................... .................... .................... .................... .................... ....................
P 6169 hCoV-19/Italy/SAR-ASSL-06/2020|EPI_ISL_614397|2020 .................... .................... .................... .................... .................... ....................
P 6170 hCoV-19/Italy/SAR-ASSL-05/2020|EPI_ISL_614398|2020 .................... .................... .................... .................... .................... ....................
P 6171 hCoV-19/Denmark/DCGC-4505/2020|EPI_ISL_615690|2020 .................... .................... .................... .................... .................... ....................
P 6172 hCoV-19/Denmark/DCGC-4506/2020|EPI_ISL_615691|2020 .................... .................... .................... .................... .................... ....................
P 6173 hCoV-19/Denmark/DCGC-4507/2020|EPI_ISL_615692|2020 .................... .................... .................... .................... .................... ....................
P 6174 hCoV-19/Denmark/DCGC-4508/2020|EPI_ISL_615693|2020 .................... .................... .................... .................... .................... ....................
P 6175 hCoV-19/Denmark/DCGC-4257/2020|EPI_ISL_615694|2020 .................... .................... .................... .................... .................... ....................
P 6176 hCoV-19/Denmark/DCGC-4258/2020|EPI_ISL_615695|2020 ........T........... .................... .................... .................... .................... ....................
P 6177 hCoV-19/Denmark/DCGC-4262/2020|EPI_ISL_615699|2020 .................... .................... .................... .................... .................... ....................
P 6178 hCoV-19/Denmark/DCGC-3281/2020|EPI_ISL_616900|2020 .................... .................... .................... .................... .................... ....................
P 6179 hCoV-19/Denmark/DCGC-3573/2020|EPI_ISL_616901|2020 .................... .................... .................... .................... .................... ....................
P 6180 hCoV-19/Denmark/DCGC-3574/2020|EPI_ISL_616902|2020 .................... .................... .................... .................... .................... ....................
P 6181 hCoV-19/Denmark/DCGC-3575/2020|EPI_ISL_616903|2020 .................... .................... .................... .................... .................... ....................
P 6182 hCoV-19/Denmark/DCGC-7056/2020|EPI_ISL_619112|2020 .................... .................... .................... .................... .................... ....................
P 6183 hCoV-19/Denmark/DCGC-7057/2020|EPI_ISL_619113|2020 .................... .................... .................... .................... .................... ....................
P 6184 hCoV-19/Denmark/DCGC-7058/2020|EPI_ISL_619114|2020 .................... .................... .................... .................... .................... ....................
P 6185 hCoV-19/Denmark/DCGC-7060/2020|EPI_ISL_619115|2020 .................... .................... .................... .................... .................... ....................
P 6186 hCoV-19/Denmark/DCGC-7061/2020|EPI_ISL_619116|2020 .................... .................... .................... .................... .................... ....................
P 6187 hCoV-19/Denmark/DCGC-7063/2020|EPI_ISL_619117|2020 .................... .................... .................... .................... .................... ....................
P 6188 hCoV-19/Denmark/DCGC-7064/2020|EPI_ISL_619118|2020 .................... .A.................. .................... .................... .................... ....................
P 6189 hCoV-19/Denmark/DCGC-7065/2020|EPI_ISL_619119|2020 .................... .N.................. .................... .................... .................... ....................
P 6190 hCoV-19/Denmark/DCGC-7066/2020|EPI_ISL_619120|2020 .................... .................... .................... .................... .................... ....................
P 6191 hCoV-19/Denmark/DCGC-7070/2020|EPI_ISL_619121|2020 .................... .................... .................... .................... .................... ....................
P 6192 hCoV-19/Denmark/DCGC-7071/2020|EPI_ISL_619122|2020 .................... .................... .................... .................... .................... ....................
P 6193 hCoV-19/Denmark/DCGC-7072/2020|EPI_ISL_619123|2020 .................... .................... .................... .................... .................... ....................
P 6194 hCoV-19/Denmark/DCGC-7073/2020|EPI_ISL_619124|2020 .................... .................... .................... .................... .................... ....................
P 6195 hCoV-19/Denmark/DCGC-7074/2020|EPI_ISL_619125|2020 .................... .................... .................... .................... .................... ....................
P 6196 hCoV-19/Denmark/DCGC-7075/2020|EPI_ISL_619126|2020 .................... .................... .................... .................... .................... ....................
P 6197 hCoV-19/Denmark/DCGC-7078/2020|EPI_ISL_619127|2020 .................... .................... .................... .................... .................... ....................
P 6198 hCoV-19/Denmark/DCGC-7079/2020|EPI_ISL_619128|2020 .................... .................... .................... .................... .................... ....................
P 6199 hCoV-19/Denmark/DCGC-7081/2020|EPI_ISL_619129|2020 .................... .................... .................... .................... .................... ....................
P 6200 hCoV-19/Denmark/DCGC-7156/2020|EPI_ISL_619173|2020 .................... .................... .................... .................... .................... ....................
P 6201 hCoV-19/Denmark/DCGC-1635/2020|EPI_ISL_619176|2020 .................... .................... .................... .................... .................... ....................
P 6202 hCoV-19/Denmark/DCGC-7162/2020|EPI_ISL_619177|2020 .................... .................... .................... .................... .................... ....................
P 6203 hCoV-19/Denmark/DCGC-7163/2020|EPI_ISL_619178|2020 .................... .................... .................... .................... .................... ....................
P 6204 hCoV-19/Denmark/DCGC-7165/2020|EPI_ISL_619179|2020 .................... .................... .................... .................... .................... ....................
P 6205 hCoV-19/Denmark/DCGC-6197/2020|EPI_ISL_620100|2020 .................... .................... .................... .................... .................... ....................
P 6206 hCoV-19/Denmark/DCGC-6198/2020|EPI_ISL_620101|2020 .................... .................... .................... .................... ....T............... ....................
P 6207 hCoV-19/Denmark/DCGC-6199/2020|EPI_ISL_620102|2020 .................... .................... .................... .................... .................... ....................
P 6208 hCoV-19/Denmark/DCGC-6200/2020|EPI_ISL_620103|2020 .................... .................... .................... .................... .................... ....................
P 6209 hCoV-19/Denmark/DCGC-6201/2020|EPI_ISL_620104|2020 .................... .................... .................... .................... .................... ....................
P 6210 hCoV-19/Denmark/DCGC-6202/2020|EPI_ISL_620105|2020 .................... .................... .................... .................... .................... ....................
P 6211 hCoV-19/Denmark/DCGC-6204/2020|EPI_ISL_620107|2020 .................... .................... .................... .................... .................... ....................
P 6212 hCoV-19/Denmark/DCGC-6206/2020|EPI_ISL_620109|2020 .................... .................... .................... .................... .................... ....................
P 6213 hCoV-19/Denmark/DCGC-6207/2020|EPI_ISL_620110|2020 .................... .................... .................... .................... .................... ....................
P 6214 hCoV-19/Denmark/DCGC-6208/2020|EPI_ISL_620111|2020 .................... .................... .................... .................... .................... ....................
P 6215 hCoV-19/Denmark/DCGC-6209/2020|EPI_ISL_620112|2020 .................... .................... .................... .................... .................... ....................
P 6216 hCoV-19/Denmark/DCGC-6210/2020|EPI_ISL_620113|2020 .................... .................... .................... .................... .................... ....................
P 6217 hCoV-19/Denmark/DCGC-6211/2020|EPI_ISL_620114|2020 .................... .................... .................... .................... .................... ....................
P 6218 hCoV-19/Denmark/DCGC-6212/2020|EPI_ISL_620115|2020 .................... .................... .................... .................... .................... ....................

| | | | | | | | | |
|---|---|---|---|---|---|---|---|---|
| P | 6219 | hCoV-19/Denmark/DCGC-5907/2020\|EPI_ISL_620172\|2020 | ···················· | ···················· | ···················· | ···················· | ···················· | ···················· |
| P | 6220 | hCoV-19/Denmark/DCGC-5908/2020\|EPI_ISL_620173\|2020 | ···················· | ···················· | ···················· | ···················· | ···················· | ···················· |
| P | 6221 | hCoV-19/Denmark/DCGC-4790/2020\|EPI_ISL_621400\|2020 | ···················· | ···················· | ···················· | ···················· | ···················· | ···················· |
| P | 6222 | hCoV-19/Denmark/DCGC-4791/2020\|EPI_ISL_621401\|2020 | ···················· | ···················· | ···················· | ···················· | ···················· | ···················· |
| P | 6223 | hCoV-19/Denmark/DCGC-4792/2020\|EPI_ISL_621402\|2020 | ···················· | ·A·················· | ···················· | ···················· | ···················· | ···················· |
| P | 6224 | hCoV-19/Denmark/DCGC-4793/2020\|EPI_ISL_621403\|2020 | ···················· | ···················· | ···················· | ···················· | ···················· | ···················· |
| P | 6225 | hCoV-19/Denmark/DCGC-4794/2020\|EPI_ISL_621404\|2020 | ···················· | ···················· | ···················· | ···················· | ···················· | ···················· |
| P | 6226 | hCoV-19/Denmark/DCGC-4795/2020\|EPI_ISL_621405\|2020 | ···················· | ···················· | ···················· | ···················· | ···················· | ···················· |
| P | 6227 | hCoV-19/Denmark/DCGC-4796/2020\|EPI_ISL_621406\|2020 | ···················· | ···················· | ···················· | ···················· | ···················· | ···················· |
| P | 6228 | hCoV-19/Denmark/DCGC-4797/2020\|EPI_ISL_621407\|2020 | ···················· | ···················· | ···················· | ···················· | ···················· | ···················· |
| P | 6229 | hCoV-19/Denmark/DCGC-4798/2020\|EPI_ISL_621408\|2020 | ···················· | ···················· | ···················· | ···················· | ···················· | ···················· |
| P | 6230 | hCoV-19/Denmark/DCGC-4799/2020\|EPI_ISL_621409\|2020 | ···················· | ···················· | ···················· | ···················· | ···················· | ···················· |
| P | 6231 | hCoV-19/Denmark/DCGC-4801/2020\|EPI_ISL_621411\|2020 | ···················· | ···················· | ···················· | ···················· | ···················· | ···················· |
| P | 6232 | hCoV-19/Denmark/DCGC-4802/2020\|EPI_ISL_621412\|2020 | ···················· | ···················· | ···················· | ···················· | ···················· | ···················· |
| P | 6233 | hCoV-19/Denmark/DCGC-4803/2020\|EPI_ISL_621413\|2020 | ···················· | ···················· | ···················· | ···················· | ···················· | ···················· |
| P | 6234 | hCoV-19/Denmark/DCGC-4804/2020\|EPI_ISL_621414\|2020 | ···················· | ···················· | ···················· | ···················· | ···················· | ···················· |
| P | 6235 | hCoV-19/Denmark/DCGC-4806/2020\|EPI_ISL_621416\|2020 | ···················· | ···················· | ···················· | ···················· | ···················· | ···················· |
| P | 6236 | hCoV-19/Denmark/DCGC-4807/2020\|EPI_ISL_621417\|2020 | ···················· | ···················· | ···················· | ···················· | ···················· | ···················· |
| P | 6237 | hCoV-19/Denmark/DCGC-4808/2020\|EPI_ISL_621418\|2020 | ···················· | ···················· | ···················· | ···················· | ···················· | ···················· |
| P | 6238 | hCoV-19/Denmark/DCGC-4809/2020\|EPI_ISL_621419\|2020 | ···················· | ···················· | ···················· | ···················· | ···················· | ···················· |
| P | 6239 | hCoV-19/Denmark/DCGC-4810/2020\|EPI_ISL_621420\|2020 | ···················· | ···················· | ···················· | ···················· | ···················· | ···················· |
| P | 6240 | hCoV-19/Denmark/DCGC-4812/2020\|EPI_ISL_621422\|2020 | ···················· | ···················· | ···················· | ···················· | ···················· | ···················· |
| P | 6241 | hCoV-19/Denmark/DCGC-4813/2020\|EPI_ISL_621423\|2020 | ···················· | ···················· | ···················· | ···················· | ···················· | ···················· |
| P | 6242 | hCoV-19/Denmark/DCGC-4814/2020\|EPI_ISL_621424\|2020 | ···················· | ···················· | ···················· | ···················· | ···················· | ···················· |
| P | 6243 | hCoV-19/Denmark/DCGC-4816/2020\|EPI_ISL_621426\|2020 | ···················· | ···················· | ···················· | ···················· | ···················· | ···················· |
| P | 6244 | hCoV-19/Denmark/DCGC-4818/2020\|EPI_ISL_621428\|2020 | ···················· | ···················· | ···················· | ···················· | ···················· | ···················· |
| P | 6245 | hCoV-19/Denmark/DCGC-4819/2020\|EPI_ISL_621429\|2020 | ···················· | ···················· | ···················· | ···················· | ···················· | ···················· |
| P | 6246 | hCoV-19/Denmark/DCGC-4820/2020\|EPI_ISL_621430\|2020 | ···················· | ···················· | ···················· | ···················· | ···················· | ···················· |
| P | 6247 | hCoV-19/Denmark/DCGC-4821/2020\|EPI_ISL_621431\|2020 | ···················· | ···················· | ···················· | ···················· | ···················· | ···················· |
| P | 6248 | hCoV-19/Denmark/DCGC-4822/2020\|EPI_ISL_621432\|2020 | ···················· | ···················· | ···················· | ···················· | ···················· | ···················· |
| P | 6249 | hCoV-19/Denmark/DCGC-4823/2020\|EPI_ISL_621433\|2020 | ···················· | ·A·················· | ···················· | ···················· | ···················· | ···················· |
| P | 6250 | hCoV-19/Denmark/DCGC-4824/2020\|EPI_ISL_621434\|2020 | ···················· | ···················· | ···················· | ···················· | ···················· | ···················· |
| P | 6251 | hCoV-19/Denmark/DCGC-4825/2020\|EPI_ISL_621435\|2020 | ···················· | ···················· | ···················· | ···················· | ···················· | ···················· |
| P | 6252 | hCoV-19/Denmark/DCGC-4826/2020\|EPI_ISL_621436\|2020 | ···················· | ···················· | ···················· | ···················· | ···················· | ···················· |
| P | 6253 | hCoV-19/Denmark/DCGC-4828/2020\|EPI_ISL_621438\|2020 | ···················· | ···················· | ···················· | ···················· | ···················· | ···················· |
| P | 6254 | hCoV-19/Denmark/DCGC-4829/2020\|EPI_ISL_621439\|2020 | ···················· | ···················· | ···················· | ···················· | ···················· | ···················· |
| P | 6255 | hCoV-19/Denmark/DCGC-4902/2020\|EPI_ISL_621440\|2020 | ···················· | ···················· | ···················· | ···················· | ···················· | ···················· |
| P | 6256 | hCoV-19/Denmark/DCGC-4903/2020\|EPI_ISL_621441\|2020 | ···················· | ···················· | ···················· | ···················· | ···················· | ···················· |
| P | 6257 | hCoV-19/Denmark/DCGC-4904/2020\|EPI_ISL_621442\|2020 | ···················· | ···················· | ···················· | ···················· | ···················· | ···················· |
| P | 6258 | hCoV-19/Denmark/DCGC-4905/2020\|EPI_ISL_621443\|2020 | ···················· | ···················· | ···················· | ···················· | ···················· | ···················· |
| P | 6259 | hCoV-19/Denmark/DCGC-4906/2020\|EPI_ISL_621444\|2020 | ···················· | ···················· | ···················· | ···················· | ···················· | ···················· |
| P | 6260 | hCoV-19/Denmark/DCGC-4907/2020\|EPI_ISL_621445\|2020 | ···················· | ···················· | ···················· | ···················· | ···················· | ···················· |
| P | 6261 | hCoV-19/Denmark/DCGC-4908/2020\|EPI_ISL_621446\|2020 | ···················· | ···················· | ···················· | ···················· | ···················· | ···················· |
| P | 6262 | hCoV-19/Denmark/DCGC-3798/2020\|EPI_ISL_622700\|2020 | ···················· | ···················· | ···················· | ···················· | ···················· | ···················· |
| P | 6263 | hCoV-19/Denmark/DCGC-3808/2020\|EPI_ISL_622702\|2020 | ···················· | ···················· | ···················· | ···················· | ···················· | ···················· |
| P | 6264 | hCoV-19/Denmark/DCGC-3813/2020\|EPI_ISL_622703\|2020 | ···················· | ···················· | ···················· | ···················· | ···················· | ···················· |
| P | 6265 | hCoV-19/Denmark/DCGC-3824/2020\|EPI_ISL_622704\|2020 | ···················· | ···················· | ···················· | ···················· | ···················· | ···················· |
| P | 6266 | hCoV-19/Denmark/DCGC-3829/2020\|EPI_ISL_622705\|2020 | ···················· | ···················· | ···················· | ···················· | ···················· | ···················· |
| P | 6267 | hCoV-19/Denmark/DCGC-3834/2020\|EPI_ISL_622706\|2020 | ···················· | ···················· | ···················· | ···················· | ···················· | ···················· |
| P | 6268 | hCoV-19/Denmark/DCGC-3838/2020\|EPI_ISL_622707\|2020 | ···················· | ···················· | ···················· | ···················· | ···················· | ···················· |
| P | 6269 | hCoV-19/Denmark/DCGC-3843/2020\|EPI_ISL_622708\|2020 | ···················· | ···················· | ···················· | ···················· | ···················· | ···················· |
| P | 6270 | hCoV-19/Denmark/DCGC-3788/2020\|EPI_ISL_622709\|2020 | ···················· | ···················· | ···················· | ···················· | ···················· | ···················· |
| P | 6271 | hCoV-19/Denmark/DCGC-3792/2020\|EPI_ISL_622710\|2020 | ···················· | ···················· | ···················· | ···················· | ···················· | ···················· |
| P | 6272 | hCoV-19/Denmark/DCGC-3799/2020\|EPI_ISL_622711\|2020 | ···················· | ···················· | ···················· | ···················· | ···················· | ···················· |
| P | 6273 | hCoV-19/Denmark/DCGC-3814/2020\|EPI_ISL_622712\|2020 | ···················· | ···················· | ···················· | ···················· | ···················· | ···················· |
| P | 6274 | hCoV-19/Denmark/DCGC-3825/2020\|EPI_ISL_622714\|2020 | ···················· | ···················· | ···················· | ···················· | ···················· | ···················· |
| P | 6275 | hCoV-19/Denmark/DCGC-3835/2020\|EPI_ISL_622715\|2020 | ···················· | ···················· | ···················· | ···················· | ···················· | ···················· |
| P | 6276 | hCoV-19/Denmark/DCGC-3839/2020\|EPI_ISL_622716\|2020 | ···················· | ···················· | ···················· | ···················· | ···················· | ···················· |
| P | 6277 | hCoV-19/Denmark/DCGC-3809/2020\|EPI_ISL_622717\|2020 | ···················· | ···················· | ···················· | ···················· | ···················· | ···················· |
| P | 6278 | hCoV-19/Denmark/DCGC-3789/2020\|EPI_ISL_622718\|2020 | ···················· | ···················· | ···················· | ···················· | ···················· | ···················· |
| P | 6279 | hCoV-19/Denmark/DCGC-3793/2020\|EPI_ISL_622719\|2020 | ···················· | ···················· | ···················· | ···················· | ···················· | ···················· |
| P | 6280 | hCoV-19/Denmark/DCGC-3805/2020\|EPI_ISL_622721\|2020 | ···················· | ···················· | ···················· | ···················· | ···················· | ···················· |
| P | 6281 | hCoV-19/Denmark/DCGC-3815/2020\|EPI_ISL_622722\|2020 | ···················· | ···················· | ···················· | ···················· | ···················· | ···················· |
| P | 6282 | hCoV-19/Denmark/DCGC-3820/2020\|EPI_ISL_622723\|2020 | ···················· | ···················· | ···················· | ···················· | ···················· | ···················· |
| P | 6283 | hCoV-19/Denmark/DCGC-1652/2020\|EPI_ISL_622724\|2020 | ···················· | ···················· | ···················· | ···················· | ···················· | ···················· |
| P | 6284 | hCoV-19/Denmark/DCGC-3830/2020\|EPI_ISL_622725\|2020 | ···················· | ···················· | ···················· | ···················· | ···················· | ···················· |
| P | 6285 | hCoV-19/Denmark/DCGC-3840/2020\|EPI_ISL_622726\|2020 | ···················· | ···················· | ···················· | ···················· | ···················· | ···················· |
| P | 6286 | hCoV-19/Denmark/DCGC-3846/2020\|EPI_ISL_622727\|2020 | ···················· | ···················· | ···················· | ···················· | ···················· | ···················· |
| P | 6287 | hCoV-19/Denmark/DCGC-3850/2020\|EPI_ISL_622728\|2020 | ···················· | ···················· | ···················· | ···················· | ···················· | ···················· |
| P | 6288 | hCoV-19/Denmark/DCGC-3844/2020\|EPI_ISL_622729\|2020 | ···················· | ···················· | ···················· | ···················· | ···················· | ···················· |
| P | 6289 | hCoV-19/Denmark/DCGC-3853/2020\|EPI_ISL_622730\|2020 | ···················· | ···················· | ···················· | ···················· | ···················· | ···················· |
| P | 6290 | hCoV-19/Denmark/DCGC-3854/2020\|EPI_ISL_622731\|2020 | ···················· | ···················· | ···················· | ···················· | ···················· | ···················· |
| P | 6291 | hCoV-19/Denmark/DCGC-3860/2020\|EPI_ISL_622732\|2020 | ···················· | ···················· | ···················· | ···················· | ···················· | ···················· |
| P | 6292 | hCoV-19/Denmark/DCGC-3861/2020\|EPI_ISL_622733\|2020 | ···················· | ···················· | ···················· | ···················· | ···················· | ···················· |
| P | 6293 | hCoV-19/Denmark/DCGC-3847/2020\|EPI_ISL_622734\|2020 | ···················· | ···················· | ···················· | ···················· | ···················· | ···················· |
| P | 6294 | hCoV-19/Denmark/DCGC-3851/2020\|EPI_ISL_622735\|2020 | ···················· | ···················· | ···················· | ···················· | ···················· | ···················· |
| P | 6295 | hCoV-19/Denmark/DCGC-3845/2020\|EPI_ISL_622736\|2020 | ···················· | ···················· | ···················· | ···················· | ···················· | ···················· |
| P | 6296 | hCoV-19/Denmark/DCGC-3856/2020\|EPI_ISL_622737\|2020 | ···················· | ···················· | ···················· | ···················· | ···················· | ···················· |
| P | 6297 | hCoV-19/Denmark/DCGC-3858/2020\|EPI_ISL_622738\|2020 | ···················· | ···················· | ···················· | ···················· | ···················· | ···················· |
| P | 6298 | hCoV-19/Denmark/DCGC-3849/2020\|EPI_ISL_622739\|2020 | ···················· | ···················· | ···················· | ···················· | ···················· | ···················· |

| | | | | | | | | |
|---|---|---|---|---|---|---|---|---|
| P | 6299 | hCoV-19/Denmark/DCGC-3864/2020\|EPI_ISL_622741\|2020 | ···················· | ···················· | ···················· | ···················· | ···················· | ···················· |
| P | 6300 | hCoV-19/Denmark/DCGC-3848/2020\|EPI_ISL_622742\|2020 | ···················· | ···················· | ···················· | ···················· | ···················· | ···················· |
| P | 6301 | hCoV-19/Denmark/DCGC-3852/2020\|EPI_ISL_622743\|2020 | ···················· | ···················· | ···················· | ···················· | ···················· | ···················· |
| P | 6302 | hCoV-19/Denmark/DCGC-3855/2020\|EPI_ISL_622744\|2020 | ···················· | ···················· | ···················· | ···················· | ···················· | ···················· |
| P | 6303 | hCoV-19/Denmark/DCGC-3857/2020\|EPI_ISL_622745\|2020 | ···················· | ···················· | ···················· | ···················· | ···················· | ···················· |
| P | 6304 | hCoV-19/Denmark/DCGC-3859/2020\|EPI_ISL_622746\|2020 | ···················· | ···················· | ···················· | ···················· | ···················· | ···················· |
| P | 6305 | hCoV-19/Denmark/DCGC-3863/2020\|EPI_ISL_622747\|2020 | ···················· | ···················· | ···················· | ···················· | ···················· | ···················· |
| P | 6306 | hCoV-19/Denmark/DCGC-4977/2020\|EPI_ISL_622748\|2020 | ···················· | ···················· | ···················· | ···················· | ···················· | ···················· |
| P | 6307 | hCoV-19/Denmark/DCGC-4991/2020\|EPI_ISL_622749\|2020 | ···················· | ···················· | ···················· | ···················· | ···················· | ···················· |
| P | 6308 | hCoV-19/Denmark/DCGC-5002/2020\|EPI_ISL_622750\|2020 | ···················· | ···················· | ···················· | ···················· | ···················· | ···················· |
| P | 6309 | hCoV-19/Denmark/DCGC-5015/2020\|EPI_ISL_622751\|2020 | ···················· | ···················· | ···················· | ···················· | ···················· | ···················· |
| P | 6310 | hCoV-19/Denmark/DCGC-5021/2020\|EPI_ISL_622752\|2020 | ···················· | ···················· | ···················· | ···················· | ···················· | ···················· |
| P | 6311 | hCoV-19/Denmark/DCGC-5028/2020\|EPI_ISL_622753\|2020 | ···················· | ···················· | ···················· | ···················· | ···················· | ···················· |
| P | 6312 | hCoV-19/Denmark/DCGC-5034/2020\|EPI_ISL_622754\|2020 | ···················· | ···················· | ···················· | ···················· | ···················· | ···················· |
| P | 6313 | hCoV-19/Denmark/DCGC-5046/2020\|EPI_ISL_622755\|2020 | ···················· | ···················· | ···················· | ···················· | ···················· | ···················· |
| P | 6314 | hCoV-19/Denmark/DCGC-4978/2020\|EPI_ISL_622756\|2020 | ···················· | ···················· | ···················· | ···················· | ···················· | ···················· |
| P | 6315 | hCoV-19/Denmark/DCGC-4984/2020\|EPI_ISL_622757\|2020 | ···················· | ···················· | ···················· | ···················· | ···················· | ···················· |
| P | 6316 | hCoV-19/Denmark/DCGC-4992/2020\|EPI_ISL_622758\|2020 | ···················· | ···················· | ···················· | ···················· | ···················· | ···················· |
| P | 6317 | hCoV-19/Denmark/DCGC-4998/2020\|EPI_ISL_622759\|2020 | ···················· | ···················· | ···················· | ···················· | ···················· | ···················· |
| P | 6318 | hCoV-19/Denmark/DCGC-5003/2020\|EPI_ISL_622760\|2020 | ···················· | ···················· | ···················· | ···················· | ···················· | ···················· |
| P | 6319 | hCoV-19/Australia/SAP523/2020\|EPI_ISL_622762\|2020- | ···················· | ···················· | ···················· | ···················· | ···················· | ···················· |
| P | 6320 | hCoV-19/Australia/SAP524/2020\|EPI_ISL_622763\|2020- | ···················· | ···················· | ···················· | ···················· | ···················· | ···················· |
| P | 6321 | hCoV-19/Australia/SAP529/2020\|EPI_ISL_622764\|2020- | ···················· | ···················· | ···················· | ···················· | ···················· | ···················· |
| P | 6322 | hCoV-19/Australia/SAP531/2020\|EPI_ISL_622765\|2020- | ···················· | ···················· | ···················· | ···················· | ···················· | ···················· |
| P | 6323 | hCoV-19/Australia/SAP532/2020\|EPI_ISL_622766\|2020- | ···················· | ···················· | ···················· | ···················· | ···················· | ···················· |
| P | 6325 | hCoV-19/New Zealand/20CV0311/2020\|EPI_ISL_622801\|2 | ···················· | ···················· | ···················· | ···················· | ···················· | ···················· |
| P | 6326 | hCoV-19/New Zealand/20CV0392/2020\|EPI_ISL_622802\|2 | ···················· | ···················· | ···················· | ···················· | ···················· | ···················· |
| P | 6327 | hCoV-19/New Zealand/20CV0397/2020\|EPI_ISL_622803\|2 | ···················· | ·A·················· | ···················· | ···················· | ···················· | ···················· |
| P | 6328 | hCoV-19/New Zealand/20CV0402/2020\|EPI_ISL_622804\|2 | ···················· | ···················· | ···················· | ···················· | ···················· | ···················· |
| P | 6329 | hCoV-19/New Zealand/20CV0404/2020\|EPI_ISL_622805\|2 | ···················· | ···················· | ···················· | ···················· | ···················· | ···················· |
| P | 6330 | hCoV-19/New Zealand/20CV0405/2020\|EPI_ISL_622806\|2 | ···················· | ···················· | ···················· | ···················· | ···················· | ···················· |
| P | 6333 | hCoV-19/New Zealand/20CV0458/2020\|EPI_ISL_622809\|2 | ···················· | ···················· | ···················· | ···················· | ···················· | ···················· |
| P | 6334 | hCoV-19/New Zealand/20CV0471/2020\|EPI_ISL_622810\|2 | ···················· | ···················· | ···················· | ···················· | ···················· | ···················· |
| P | 6335 | hCoV-19/England/ALDP-AA0B8F/2020\|EPI_ISL_625010\|20 | ···················· | ···················· | ···················· | ···················· | ···················· | ···················· |
| P | 6336 | hCoV-19/England/ALDP-AA16D8/2020\|EPI_ISL_625011\|20 | ···················· | ·A·················· | ···················· | ···················· | ···················· | ··A················· |
| P | 6337 | hCoV-19/England/ALDP-AA5461/2020\|EPI_ISL_625012\|20 | ···················· | ···················· | ···················· | ···················· | ···················· | ···················· |
| P | 6338 | hCoV-19/England/ALDP-AA35F7/2020\|EPI_ISL_625014\|20 | ···················· | ·A·················· | ···················· | ···················· | ···················· | ···················· |
| P | 6339 | hCoV-19/England/ALDP-AA5319/2020\|EPI_ISL_625015\|20 | ···················· | ···················· | ···················· | ···················· | ···················· | ···················· |
| P | 6340 | hCoV-19/England/ALDP-AA0BAD/2020\|EPI_ISL_625016\|20 | ···················· | ···················· | ···················· | ···················· | ···················· | ···················· |
| P | 6341 | hCoV-19/England/ALDP-AA4505/2020\|EPI_ISL_625019\|20 | ···················· | ···················· | ···················· | ···················· | ···················· | ···················· |
| P | 6342 | hCoV-19/England/ALDP-AA167E/2020\|EPI_ISL_625020\|20 | ···················· | ···················· | ···················· | ···················· | ···················· | ···················· |
| P | 6343 | hCoV-19/England/ALDP-AA0C04/2020\|EPI_ISL_625021\|20 | ···················· | ···················· | ···················· | ···················· | ···················· | ···················· |
| P | 6344 | hCoV-19/England/ALDP-AA1E1C/2020\|EPI_ISL_625022\|20 | ···················· | ···················· | ···················· | ···················· | ···················· | ···················· |
| P | 6345 | hCoV-19/England/ALDP-AA38C1/2020\|EPI_ISL_625023\|20 | ···················· | ···················· | ···················· | ···················· | ···················· | ···················· |
| P | 6346 | hCoV-19/Wales/ALDP-AA4963/2020\|EPI_ISL_625024\|2020 | ···················· | ···················· | ···················· | ···················· | ······T············· | ···················· |
| P | 6347 | hCoV-19/England/ALDP-AA54F8/2020\|EPI_ISL_625025\|20 | ···················· | ···················· | ···················· | ···················· | ···················· | ···················· |
| P | 6348 | hCoV-19/England/ALDP-AA3436/2020\|EPI_ISL_625027\|20 | ···················· | ·A·················· | ···················· | ···················· | ···T················ | ···················· |
| P | 6349 | hCoV-19/England/ALDP-AA0A46/2020\|EPI_ISL_625028\|20 | ···················· | ···················· | ···················· | ···················· | ···················· | ···················· |
| P | 6350 | hCoV-19/England/ALDP-AA08F1/2020\|EPI_ISL_625029\|20 | ···················· | ···················· | ···················· | ···················· | ···················· | ···················· |
| P | 6351 | hCoV-19/England/ALDP-AA2D2D/2020\|EPI_ISL_625030\|20 | ···················· | ···················· | ···················· | ···················· | ···················· | ···················· |
| P | 6352 | hCoV-19/England/ALDP-AA44F9/2020\|EPI_ISL_625031\|20 | ···················· | ···················· | ···················· | ···················· | ···················· | ···················· |
| P | 6353 | hCoV-19/England/ALDP-AA3EFC/2020\|EPI_ISL_625032\|20 | ···················· | ···················· | ···················· | ···················· | ···················· | ···················· |
| P | 6354 | hCoV-19/England/ALDP-AA5407/2020\|EPI_ISL_625033\|20 | ···················· | ·A·················· | ···················· | ···················· | ···················· | ··A················· |
| P | 6355 | hCoV-19/England/ALDP-AA1131/2020\|EPI_ISL_625034\|20 | ···················· | ···················· | ···················· | ···················· | ···················· | ···················· |
| P | 6356 | hCoV-19/England/ALDP-AA33FD/2020\|EPI_ISL_625035\|20 | ···················· | ···················· | ···················· | ···················· | ···················· | ···················· |
| P | 6357 | hCoV-19/England/ALDP-AA2279/2020\|EPI_ISL_625036\|20 | ···················· | ···················· | ···················· | ···················· | ···················· | ···················· |
| P | 6358 | hCoV-19/England/ALDP-AA36A9/2020\|EPI_ISL_625037\|20 | ···················· | ···················· | ···················· | ···················· | ···················· | ···················· |
| P | 6359 | hCoV-19/England/ALDP-AA4514/2020\|EPI_ISL_625038\|20 | ···················· | ···················· | ···················· | ···················· | ···················· | ···················· |
| P | 6360 | hCoV-19/England/ALDP-AA325A/2020\|EPI_ISL_625039\|20 | ···················· | ···················· | ···················· | ···················· | ···················· | ···················· |
| P | 6361 | hCoV-19/England/ALDP-AA556E/2020\|EPI_ISL_625040\|20 | ···················· | ···················· | ···················· | ···················· | ···················· | ···················· |
| P | 6362 | hCoV-19/England/ALDP-AA3F08/2020\|EPI_ISL_625041\|20 | ···················· | ···················· | ···················· | ···················· | ···················· | ···················· |
| P | 6363 | hCoV-19/England/ALDP-AA559B/2020\|EPI_ISL_625042\|20 | ···················· | ···················· | ···················· | ···················· | ···················· | ···················· |
| P | 6364 | hCoV-19/England/ALDP-AA2112/2020\|EPI_ISL_625043\|20 | ···················· | ···················· | ···················· | ···················· | ···················· | ···················· |
| P | 6365 | hCoV-19/England/ALDP-AA5470/2020\|EPI_ISL_625044\|20 | ···················· | ···················· | ···················· | ···················· | ···················· | ···················· |
| P | 6366 | hCoV-19/England/ALDP-AA2D1E/2020\|EPI_ISL_625045\|20 | ···················· | ···················· | ···················· | ···················· | ···················· | ···················· |
| P | 6367 | hCoV-19/England/ALDP-AA2297/2020\|EPI_ISL_625046\|20 | ···················· | ···················· | ···················· | ···················· | ···················· | ···················· |
| P | 6368 | hCoV-19/England/ALDP-AA2C8A/2020\|EPI_ISL_625047\|20 | ···················· | ···················· | ···················· | ···················· | ···················· | ···················· |
| P | 6369 | hCoV-19/England/ALDP-AA33EE/2020\|EPI_ISL_625048\|20 | ···················· | ···················· | ···················· | ···················· | ···················· | ···················· |
| P | 6370 | hCoV-19/England/ALDP-AA178A/2020\|EPI_ISL_625050\|20 | ···················· | ···················· | ···················· | ···················· | ···················· | ···················· |
| P | 6371 | hCoV-19/England/ALDP-AA2C4E/2020\|EPI_ISL_625051\|20 | ···················· | ···················· | ···················· | ···················· | ···················· | ···················· |
| P | 6372 | hCoV-19/England/ALDP-AA219A/2020\|EPI_ISL_625052\|20 | ···················· | ···················· | ···················· | ···················· | ···················· | ···················· |
| P | 6373 | hCoV-19/England/ALDP-AA0CAA/2020\|EPI_ISL_625053\|20 | ···················· | ···················· | ···················· | ···················· | ···················· | ···················· |
| P | 6374 | hCoV-19/England/ALDP-AA35E8/2020\|EPI_ISL_625054\|20 | ···················· | ···················· | ···················· | ···················· | ······T············· | ···················· |
| P | 6375 | hCoV-19/England/ALDP-AA2C7B/2020\|EPI_ISL_625055\|20 | ···················· | ·A·················· | ···················· | ···················· | ···················· | ··A················· |
| P | 6376 | hCoV-19/England/ALDP-AA35D9/2020\|EPI_ISL_625056\|20 | ···················· | ···················· | ···················· | ···················· | ···················· | ···················· |
| P | 6377 | hCoV-19/England/ALDP-AA52B2/2020\|EPI_ISL_625057\|20 | ···················· | ···················· | ···················· | ···················· | ···················· | ···················· |
| P | 6378 | hCoV-19/England/ALDP-AA53BF/2020\|EPI_ISL_625058\|20 | ···················· | ···················· | ···················· | ···················· | ···················· | ···················· |
| P | 6379 | hCoV-19/England/ALDP-AA3427/2020\|EPI_ISL_625059\|20 | ···················· | ···················· | ···················· | ···················· | ···················· | ···················· |
| P | 6380 | hCoV-19/England/ALDP-AA457E/2020\|EPI_ISL_625067\|20 | ···················· | ···················· | ···················· | ···················· | ···················· | ···················· |
| P | 6381 | hCoV-19/England/ALDP-AA55C8/2020\|EPI_ISL_625068\|20 | ···················· | ···················· | ···················· | ···················· | ···················· | ···················· |

| | | | | | | | | |
|---|---|---|---|---|---|---|---|---|
| P | 6382 | hCoV-19/England/ALDP-AA0ACE/2020\|EPI_ISL_625070\|20 | ···················· | ···················· | ···················· | ···················· | ···················· | ···················· |
| P | 6383 | hCoV-19/England/ALDP-AA0B52/2020\|EPI_ISL_625071\|20 | ···················· | ·A·················· | ···················· | ···················· | ···················· | ···················· |
| P | 6384 | hCoV-19/England/ALDP-AA1702/2020\|EPI_ISL_625072\|20 | ···················· | ···················· | ···················· | ···················· | ······T············· | ···················· |
| P | 6385 | hCoV-19/England/ALDP-AA32A5/2020\|EPI_ISL_625073\|20 | ···················· | ···················· | ···················· | ···················· | ···················· | ···················· |
| P | 6386 | hCoV-19/England/ALDP-AA1DC4/2020\|EPI_ISL_625074\|20 | ···················· | ···················· | ···················· | ···················· | ···················· | ···················· |
| P | 6387 | hCoV-19/England/ALDP-AA21F4/2020\|EPI_ISL_625075\|20 | ···················· | ···················· | ···················· | ···················· | ···················· | ···················· |
| P | 6388 | hCoV-19/England/ALDP-AA49DC/2020\|EPI_ISL_625076\|20 | ···················· | ···················· | ···················· | ···················· | ···················· | ···················· |
| P | 6389 | hCoV-19/England/ALDP-AA5610/2020\|EPI_ISL_625077\|20 | ···················· | ···················· | ···················· | ···················· | ···················· | ···················· |
| P | 6390 | hCoV-19/England/ALDP-AA0D01/2020\|EPI_ISL_625078\|20 | ···················· | ···················· | ···················· | ···················· | ···················· | ···················· |
| P | 6391 | hCoV-19/England/ALDP-AA3EB0/2020\|EPI_ISL_625079\|20 | ···················· | ···················· | ···················· | ···················· | ···················· | ···················· |
| P | 6392 | hCoV-19/England/ALDP-AA3EA1/2020\|EPI_ISL_625080\|20 | ···················· | ···················· | ···················· | ···················· | ···················· | ···················· |
| P | 6393 | hCoV-19/England/ALDP-AA54AD/2020\|EPI_ISL_625081\|20 | ···················· | ···················· | ···················· | ···················· | ···················· | ···················· |
| P | 6394 | hCoV-19/England/ALDP-AA1887/2020\|EPI_ISL_625082\|20 | ···················· | ···················· | ···················· | ···················· | ···················· | ···················· |
| P | 6395 | hCoV-19/England/ALDP-AA4523/2020\|EPI_ISL_625083\|20 | ···················· | ···················· | ···················· | ···················· | ···················· | ···················· |
| P | 6396 | hCoV-19/England/ALDP-AA1DA6/2020\|EPI_ISL_625084\|20 | ···················· | ···················· | ···················· | ···················· | ···················· | ···················· |
| P | 6397 | hCoV-19/England/ALDP-AA555F/2020\|EPI_ISL_625085\|20 | ············T······· | ···················· | ···················· | ···················· | ···················· | ···················· |
| P | 6398 | hCoV-19/England/ALDP-AA566B/2020\|EPI_ISL_625086\|20 | ···················· | ···················· | ···················· | ···················· | ···················· | ···················· |
| P | 6399 | hCoV-19/England/ALDP-AA2D0F/2020\|EPI_ISL_625087\|20 | ···················· | ···················· | ···················· | ···················· | ···················· | ···················· |
| P | 6400 | hCoV-19/England/ALDP-AA20CA/2020\|EPI_ISL_625088\|20 | ···················· | ···················· | ···················· | ···················· | ···················· | ···················· |
| P | 6401 | hCoV-19/Denmark/DCGC-4330/2020\|EPI_ISL_615530\|2020 | ···················· | ···················· | ···················· | ···················· | ···················· | ···················· |
| P | 6402 | hCoV-19/Denmark/DCGC-4331/2020\|EPI_ISL_615531\|2020 | ···················· | ···················· | ···················· | ···················· | ···················· | ···················· |
| P | 6403 | hCoV-19/Denmark/DCGC-4332/2020\|EPI_ISL_615532\|2020 | ···················· | ···················· | ···················· | ···················· | ···················· | ···················· |
| P | 6404 | hCoV-19/Denmark/DCGC-4333/2020\|EPI_ISL_615533\|2020 | ···················· | ···················· | ···················· | ···················· | ···················· | ···················· |
| P | 6405 | hCoV-19/Denmark/DCGC-4334/2020\|EPI_ISL_615534\|2020 | ···················· | ···················· | ···················· | ···················· | ···················· | ···················· |
| P | 6406 | hCoV-19/Denmark/DCGC-4335/2020\|EPI_ISL_615535\|2020 | ···················· | ···················· | ···················· | ···················· | ···················· | ···················· |
| P | 6407 | hCoV-19/Denmark/DCGC-4336/2020\|EPI_ISL_615536\|2020 | ···················· | ···················· | ···················· | ···················· | ···················· | ···················· |
| P | 6408 | hCoV-19/Denmark/DCGC-4337/2020\|EPI_ISL_615537\|2020 | ···················· | ···················· | ···················· | ···················· | ···················· | ···················· |
| P | 6409 | hCoV-19/Denmark/DCGC-6313/2020\|EPI_ISL_615538\|2020 | ···················· | ···················· | ···················· | ···················· | ···················· | ···················· |
| P | 6410 | hCoV-19/Denmark/DCGC-4338/2020\|EPI_ISL_615539\|2020 | ···················· | ···················· | ···················· | ···················· | ···················· | ···················· |
| P | 6411 | hCoV-19/Denmark/DCGC-6314/2020\|EPI_ISL_615540\|2020 | ···················· | ···················· | ···················· | ···················· | ···················· | ···················· |
| P | 6412 | hCoV-19/Denmark/DCGC-4339/2020\|EPI_ISL_615541\|2020 | ···················· | ···················· | ···················· | ···················· | ···················· | ···················· |
| P | 6413 | hCoV-19/Denmark/DCGC-4340/2020\|EPI_ISL_615542\|2020 | ···················· | ···················· | ···················· | ···················· | ···················· | ···················· |
| P | 6414 | hCoV-19/Denmark/DCGC-4342/2020\|EPI_ISL_615544\|2020 | ···················· | ···················· | ···················· | ···················· | ···················· | ···················· |
| P | 6415 | hCoV-19/Denmark/DCGC-4343/2020\|EPI_ISL_615545\|2020 | ···················· | ···················· | ···················· | ···················· | ···················· | ···················· |
| P | 6416 | hCoV-19/Denmark/DCGC-4344/2020\|EPI_ISL_615546\|2020 | ···················· | ···················· | ···················· | ···················· | ···················· | ···················· |
| P | 6417 | hCoV-19/Denmark/DCGC-4345/2020\|EPI_ISL_615547\|2020 | ···················· | ···················· | ···················· | ···················· | ···················· | ···················· |
| P | 6418 | hCoV-19/Denmark/DCGC-4346/2020\|EPI_ISL_615548\|2020 | ···················· | ···················· | ···················· | ···················· | ···················· | ···················· |
| P | 6419 | hCoV-19/Denmark/DCGC-4347/2020\|EPI_ISL_615549\|2020 | ···················· | ···················· | ···················· | ···················· | ···················· | ···················· |
| P | 6420 | hCoV-19/Denmark/DCGC-4348/2020\|EPI_ISL_615550\|2020 | ···················· | ···················· | ···················· | ···················· | ···················· | ···················· |
| P | 6421 | hCoV-19/Denmark/DCGC-4350/2020\|EPI_ISL_615552\|2020 | ···················· | ···················· | ···················· | ···················· | ···················· | ···················· |
| P | 6422 | hCoV-19/Denmark/DCGC-4351/2020\|EPI_ISL_615553\|2020 | ···················· | ···················· | ···················· | ···················· | ···················· | ···················· |
| P | 6423 | hCoV-19/Denmark/DCGC-4352/2020\|EPI_ISL_615554\|2020 | ···················· | ···················· | ···················· | ···················· | ···················· | ···················· |
| P | 6424 | hCoV-19/Denmark/DCGC-4353/2020\|EPI_ISL_615555\|2020 | ···················· | ···················· | ···················· | ···················· | ···················· | ···················· |
| P | 6425 | hCoV-19/Denmark/DCGC-4354/2020\|EPI_ISL_615556\|2020 | ········T··········· | ···················· | ···················· | ···················· | ···················· | ···················· |
| P | 6426 | hCoV-19/Denmark/DCGC-4355/2020\|EPI_ISL_615557\|2020 | ···················· | ···················· | ···················· | ···················· | ···················· | ···················· |
| P | 6427 | hCoV-19/Denmark/DCGC-4356/2020\|EPI_ISL_615558\|2020 | ···················· | ···················· | ···················· | ···················· | ···················· | ···················· |
| P | 6428 | hCoV-19/Denmark/DCGC-4362/2020\|EPI_ISL_615565\|2020 | ···················· | ···················· | ···················· | ···················· | ···················· | ···················· |
| P | 6429 | hCoV-19/Denmark/DCGC-4363/2020\|EPI_ISL_615566\|2020 | ···················· | ···················· | ···················· | ···················· | ···················· | ···················· |
| P | 6430 | hCoV-19/Denmark/DCGC-4366/2020\|EPI_ISL_615569\|2020 | ···················· | ···················· | ···················· | ···················· | ···················· | ···················· |
| P | 6431 | hCoV-19/Denmark/DCGC-3882/2020\|EPI_ISL_615600\|2020 | ···················· | ···················· | ···················· | ···················· | ···················· | ···················· |
| P | 6432 | hCoV-19/Denmark/DCGC-6317/2020\|EPI_ISL_615601\|2020 | ···················· | ···················· | ···················· | ···················· | ···················· | ···················· |
| P | 6433 | hCoV-19/Denmark/DCGC-3883/2020\|EPI_ISL_615602\|2020 | ···················· | ···················· | ···················· | ···················· | ···················· | ···················· |
| P | 6434 | hCoV-19/Denmark/DCGC-3884/2020\|EPI_ISL_615603\|2020 | ···················· | ···················· | ···················· | ···················· | ···················· | ···················· |
| P | 6435 | hCoV-19/Denmark/DCGC-3885/2020\|EPI_ISL_615604\|2020 | ···················· | ···················· | ···················· | ···················· | ···················· | ···················· |
| P | 6436 | hCoV-19/Denmark/DCGC-3886/2020\|EPI_ISL_615605\|2020 | ···················· | ···················· | ···················· | ···················· | ···················· | ···················· |
| P | 6437 | hCoV-19/Denmark/DCGC-3080/2020\|EPI_ISL_616860\|2020 | ···················· | ···················· | ···················· | ···················· | ···················· | ···················· |
| P | 6438 | hCoV-19/Denmark/DCGC-3081/2020\|EPI_ISL_616861\|2020 | ···················· | ···················· | ···················· | ···················· | ···················· | ···················· |
| P | 6439 | hCoV-19/Denmark/DCGC-3083/2020\|EPI_ISL_616863\|2020 | ···················· | ···················· | ···················· | ···················· | ···················· | ···················· |
| P | 6440 | hCoV-19/Denmark/DCGC-3085/2020\|EPI_ISL_616865\|2020 | ········T··········· | ···················· | ···················· | ···················· | ···················· | ···················· |
| P | 6441 | hCoV-19/Denmark/DCGC-3086/2020\|EPI_ISL_616866\|2020 | ···················· | ···················· | ···················· | ···················· | ···················· | ···················· |
| P | 6442 | hCoV-19/Denmark/DCGC-3087/2020\|EPI_ISL_616867\|2020 | ···················· | ·A·················· | ···················· | ···················· | ···················· | ···················· |
| P | 6443 | hCoV-19/Denmark/DCGC-3088/2020\|EPI_ISL_616868\|2020 | ···················· | ···················· | ···················· | ···················· | ···················· | ···················· |
| P | 6444 | hCoV-19/Denmark/DCGC-3089/2020\|EPI_ISL_616869\|2020 | ········T··········· | ···················· | ···················· | ···················· | ···················· | ···················· |
| P | 6445 | hCoV-19/Denmark/DCGC-3090/2020\|EPI_ISL_616870\|2020 | ···················· | ···················· | ···················· | ···················· | ···················· | ···················· |
| P | 6446 | hCoV-19/Denmark/DCGC-3091/2020\|EPI_ISL_616871\|2020 | ···················· | ···················· | ···················· | ···················· | ···················· | ···················· |
| P | 6447 | hCoV-19/Denmark/DCGC-3092/2020\|EPI_ISL_616872\|2020 | ···················· | ···················· | ···················· | ···················· | ···················· | ···················· |
| P | 6448 | hCoV-19/Denmark/DCGC-3093/2020\|EPI_ISL_616873\|2020 | ···················· | ···················· | ···················· | ···················· | ···················· | ···················· |
| P | 6449 | hCoV-19/Denmark/DCGC-3095/2020\|EPI_ISL_616875\|2020 | ···················· | ···················· | ···················· | ···················· | ···················· | ···················· |
| P | 6450 | hCoV-19/Denmark/DCGC-3244/2020\|EPI_ISL_616876\|2020 | ···················· | ···················· | ···················· | ···················· | ···················· | ···················· |
| P | 6451 | hCoV-19/Denmark/DCGC-3245/2020\|EPI_ISL_616877\|2020 | ········T··········· | ···················· | ···················· | ···················· | ···················· | ···················· |
| P | 6452 | hCoV-19/Denmark/DCGC-3246/2020\|EPI_ISL_616878\|2020 | ···················· | ···················· | ···················· | ···················· | ···················· | ···················· |
| P | 6453 | hCoV-19/Denmark/DCGC-3219/2020\|EPI_ISL_616879\|2020 | ···················· | ···················· | ···················· | ···················· | ···················· | ···················· |
| P | 6454 | hCoV-19/Denmark/DCGC-3220/2020\|EPI_ISL_616880\|2020 | ···················· | ···················· | ···················· | ···················· | ···················· | ···················· |
| P | 6455 | hCoV-19/Denmark/DCGC-3221/2020\|EPI_ISL_616881\|2020 | ···················· | ···················· | ···················· | ···················· | ···················· | ···················· |
| P | 6456 | hCoV-19/Denmark/DCGC-3222/2020\|EPI_ISL_616882\|2020 | ···················· | ···················· | ···················· | ···················· | ···················· | ···················· |
| P | 6457 | hCoV-19/Denmark/DCGC-3223/2020\|EPI_ISL_616883\|2020 | ···················· | ···················· | ···················· | ···················· | ···················· | ···················· |
| P | 6458 | hCoV-19/Denmark/DCGC-3224/2020\|EPI_ISL_616884\|2020 | ···················· | ···················· | ···················· | ···················· | ···················· | ···················· |
| P | 6459 | hCoV-19/Denmark/DCGC-3225/2020\|EPI_ISL_616885\|2020 | ···················· | ···················· | ···················· | ···················· | ···················· | ···················· |
| P | 6460 | hCoV-19/Denmark/DCGC-3226/2020\|EPI_ISL_616886\|2020 | ···················· | ···················· | ···················· | ···················· | ···················· | ···················· |
| P | 6461 | hCoV-19/Denmark/DCGC-3227/2020\|EPI_ISL_616887\|2020 | ········T··········· | ···················· | ···················· | ···················· | ···················· | ···················· |

| | | | | | | | | |
|---|---|---|---|---|---|---|---|---|
| P | 6462 | hCoV-19/Denmark/DCGC-3228/2020\|EPI_ISL_616888\|2020 | .................... | .................... | ..................... | .................... | .................... | .................... |
| P | 6463 | hCoV-19/Denmark/DCGC-3229/2020\|EPI_ISL_616889\|2020 | .................... | .................... | ..................... | .................... | .................... | .................... |
| P | 6464 | hCoV-19/Denmark/DCGC-3232/2020\|EPI_ISL_616892\|2020 | .................... | .................... | ..................... | .................... | .................... | .................... |
| P | 6465 | hCoV-19/Denmark/DCGC-3236/2020\|EPI_ISL_616896\|2020 | .................... | .................... | ..................... | .................... | .................... | .................... |
| P | 6466 | hCoV-19/Denmark/DCGC-3237/2020\|EPI_ISL_616897\|2020 | .................... | .................... | ..................... | .................... | .................... | .................... |
| P | 6467 | hCoV-19/Denmark/DCGC-3238/2020\|EPI_ISL_616898\|2020 | .................... | .................... | ..................... | .................... | .................... | .................... |
| P | 6468 | hCoV-19/Denmark/DCGC-3239/2020\|EPI_ISL_616899\|2020 | .................... | .................... | ..................... | .................... | .................... | .................... |
| P | 6469 | hCoV-19/Denmark/DCGC-3577/2020\|EPI_ISL_616905\|2020 | .................... | .................... | ..................... | .................... | .................... | .................... |
| P | 6470 | hCoV-19/Denmark/DCGC-7084/2020\|EPI_ISL_619130\|2020 | .................... | .................... | ..................... | .................... | .................... | .................... |
| P | 6471 | hCoV-19/Denmark/DCGC-7088/2020\|EPI_ISL_619131\|2020 | .................... | .................... | ..................... | .................... | .................... | .................... |
| P | 6472 | hCoV-19/Denmark/DCGC-7090/2020\|EPI_ISL_619132\|2020 | .................... | .................... | ..................... | .................... | .................... | .................... |
| P | 6473 | hCoV-19/Denmark/DCGC-1642/2020\|EPI_ISL_619133\|2020 | .................... | .................... | ..................... | .................... | .................... | .................... |
| P | 6474 | hCoV-19/Denmark/DCGC-7091/2020\|EPI_ISL_619134\|2020 | .................... | .................... | ..................... | .................... | .................... | .................... |
| P | 6475 | hCoV-19/Denmark/DCGC-7093/2020\|EPI_ISL_619135\|2020 | .................... | .................... | ..................... | .................... | .................... | .................... |
| P | 6476 | hCoV-19/Denmark/DCGC-7094/2020\|EPI_ISL_619136\|2020 | .................... | .................... | ..................... | .................... | .................... | .................... |
| P | 6477 | hCoV-19/Denmark/DCGC-7095/2020\|EPI_ISL_619137\|2020 | .................... | .................... | ..................... | .................... | ....T............... | .................... |
| P | 6478 | hCoV-19/Denmark/DCGC-7096/2020\|EPI_ISL_619138\|2020 | .................... | .................... | ..................... | ............A....... | .................... | .................... |
| P | 6479 | hCoV-19/Denmark/DCGC-7098/2020\|EPI_ISL_619139\|2020 | .................... | .A.................. | ..................... | .................... | .................... | ...A................ |
| P | 6480 | hCoV-19/Denmark/DCGC-7100/2020\|EPI_ISL_619140\|2020 | .................... | .................... | ..................... | .................... | .................... | .................... |
| P | 6481 | hCoV-19/Denmark/DCGC-1641/2020\|EPI_ISL_619141\|2020 | .................... | .................... | ..................... | .................... | .................... | .................... |
| P | 6482 | hCoV-19/Denmark/DCGC-7101/2020\|EPI_ISL_619142\|2020 | .................... | .................... | ..................... | .................... | .................... | .................... |
| P | 6483 | hCoV-19/Denmark/DCGC-7102/2020\|EPI_ISL_619143\|2020 | .................... | .................... | ..................... | .................... | .................... | .................... |
| P | 6484 | hCoV-19/Denmark/DCGC-7105/2020\|EPI_ISL_619144\|2020 | .................... | .................... | ..................... | .................... | .................... | .................... |
| P | 6485 | hCoV-19/Denmark/DCGC-1640/2020\|EPI_ISL_619145\|2020 | .................... | .................... | ..................... | .................... | .................... | .................... |
| P | 6486 | hCoV-19/Denmark/DCGC-7110/2020\|EPI_ISL_619146\|2020 | .................... | .................... | ..................... | .................... | .................... | .................... |
| P | 6487 | hCoV-19/Denmark/DCGC-7111/2020\|EPI_ISL_619147\|2020 | .................... | .................... | ..................... | .................... | .................... | .................... |
| P | 6488 | hCoV-19/Denmark/DCGC-7112/2020\|EPI_ISL_619148\|2020 | .................... | .................... | ..................... | .................... | .................... | .................... |
| P | 6489 | hCoV-19/Denmark/DCGC-7113/2020\|EPI_ISL_619149\|2020 | .................... | .................... | ..................... | .................... | .................... | .................... |
| P | 6490 | hCoV-19/Denmark/DCGC-7115/2020\|EPI_ISL_619150\|2020 | .................... | .................... | ..................... | .................... | .................... | .................... |
| P | 6491 | hCoV-19/Denmark/DCGC-1639/2020\|EPI_ISL_619151\|2020 | .................... | .................... | ..................... | .................... | .................... | .................... |
| P | 6492 | hCoV-19/Denmark/DCGC-7119/2020\|EPI_ISL_619152\|2020 | .................... | .A.................. | ..................... | .................... | .................... | ...A................ |
| P | 6493 | hCoV-19/Denmark/DCGC-7123/2020\|EPI_ISL_619153\|2020 | .................... | .................... | ..................... | .................... | .................... | .................... |
| P | 6494 | hCoV-19/Denmark/DCGC-7124/2020\|EPI_ISL_619154\|2020 | .................... | .................... | ..................... | .................... | .................... | .................... |
| P | 6495 | hCoV-19/Denmark/DCGC-7125/2020\|EPI_ISL_619155\|2020 | .................... | .A.................. | ..................... | .................... | .................... | .................... |
| P | 6496 | hCoV-19/Denmark/DCGC-7126/2020\|EPI_ISL_619156\|2020 | .................... | .A.................. | ..................... | .................... | .................... | .................... |
| P | 6497 | hCoV-19/Denmark/DCGC-7129/2020\|EPI_ISL_619157\|2020 | .................... | .................... | ..................... | .................... | .................... | .................... |
| P | 6498 | hCoV-19/Denmark/DCGC-7131/2020\|EPI_ISL_619158\|2020 | .................... | .................... | ..................... | .................... | .................... | .................... |
| P | 6499 | hCoV-19/Denmark/DCGC-7133/2020\|EPI_ISL_619159\|2020 | .................... | .................... | ..................... | .................... | .................... | .................... |
| P | 6500 | hCoV-19/Denmark/DCGC-7136/2020\|EPI_ISL_619160\|2020 | .................... | .................... | ..................... | .................... | .................... | .................... |
| P | 6501 | hCoV-19/Denmark/DCGC-7140/2020\|EPI_ISL_619161\|2020 | .................... | .................... | ..................... | .................... | .................... | .................... |
| P | 6502 | hCoV-19/Denmark/DCGC-7141/2020\|EPI_ISL_619162\|2020 | .................... | .................... | ..................... | .................... | .................... | .................... |
| P | 6503 | hCoV-19/Denmark/DCGC-1637/2020\|EPI_ISL_619163\|2020 | .................... | .................... | ..................... | .................... | .................... | .................... |
| P | 6504 | hCoV-19/Denmark/DCGC-7144/2020\|EPI_ISL_619164\|2020 | .................... | .................... | ..................... | .................... | .................... | .................... |
| P | 6505 | hCoV-19/Denmark/DCGC-7145/2020\|EPI_ISL_619165\|2020 | .................... | .................... | ..................... | .................... | .................... | .................... |
| P | 6506 | hCoV-19/Denmark/DCGC-7146/2020\|EPI_ISL_619166\|2020 | .................... | .................... | ..................... | .................... | .................... | .................... |
| P | 6507 | hCoV-19/Denmark/DCGC-7147/2020\|EPI_ISL_619167\|2020 | .................... | .................... | ..................... | .................... | .................... | .................... |
| P | 6508 | hCoV-19/Denmark/DCGC-7149/2020\|EPI_ISL_619168\|2020 | .................... | .................... | ..................... | .................... | .................... | .................... |
| P | 6509 | hCoV-19/Denmark/DCGC-7151/2020\|EPI_ISL_619169\|2020 | .................... | .................... | ..................... | .................... | .................... | .................... |
| P | 6510 | hCoV-19/Denmark/DCGC-1636/2020\|EPI_ISL_619170\|2020 | .................... | .A.................. | ..................... | .................... | .................... | .................... |
| P | 6511 | hCoV-19/Denmark/DCGC-7152/2020\|EPI_ISL_619171\|2020 | .................... | .................... | ..................... | .................... | .................... | .................... |
| P | 6512 | hCoV-19/Denmark/DCGC-7153/2020\|EPI_ISL_619172\|2020 | .................... | .................... | ..................... | .................... | .................... | .................... |
| P | 6513 | hCoV-19/Denmark/DCGC-7158/2020\|EPI_ISL_619174\|2020 | .................... | .................... | ..................... | .................... | .................... | .................... |
| P | 6514 | hCoV-19/Denmark/DCGC-7159/2020\|EPI_ISL_619175\|2020 | .................... | .................... | ..................... | .................... | .................... | .................... |
| P | 6515 | hCoV-19/Denmark/DCGC-7166/2020\|EPI_ISL_619180\|2020 | .................... | .................... | ..................... | .................... | .................... | .................... |
| P | 6516 | hCoV-19/Denmark/DCGC-7170/2020\|EPI_ISL_619181\|2020 | .................... | .................... | ..................... | .................... | .................... | .................... |
| P | 6517 | hCoV-19/Denmark/DCGC-7171/2020\|EPI_ISL_619182\|2020 | .................... | .................... | ..................... | .................... | .................... | .................... |
| P | 6518 | hCoV-19/Denmark/DCGC-7172/2020\|EPI_ISL_619183\|2020 | .................... | .................... | ..................... | .................... | .................... | .................... |
| P | 6519 | hCoV-19/Denmark/DCGC-7175/2020\|EPI_ISL_619184\|2020 | .................... | .................... | ..................... | .................... | .................... | .................... |
| P | 6520 | hCoV-19/Denmark/DCGC-1633/2020\|EPI_ISL_619185\|2020 | .................... | .................... | ..................... | .................... | .................... | .................... |
| P | 6521 | hCoV-19/Denmark/DCGC-7181/2020\|EPI_ISL_619186\|2020 | .................... | .................... | ..................... | .................... | .................... | .................... |
| P | 6522 | hCoV-19/Denmark/DCGC-7182/2020\|EPI_ISL_619187\|2020 | .................... | .................... | ..................... | .................... | .................... | .................... |
| P | 6523 | hCoV-19/Denmark/DCGC-7187/2020\|EPI_ISL_619188\|2020 | .................... | .................... | ..................... | .................... | .................... | .................... |
| P | 6524 | hCoV-19/Denmark/DCGC-7188/2020\|EPI_ISL_619189\|2020 | .................... | .................... | ..................... | .................... | .................... | .................... |
| P | 6525 | hCoV-19/Denmark/DCGC-7189/2020\|EPI_ISL_619190\|2020 | .................... | .................... | ..................... | .................... | .................... | .................... |
| P | 6526 | hCoV-19/Denmark/DCGC-7191/2020\|EPI_ISL_619191\|2020 | .................... | .................... | ..................... | .................... | .................... | .................... |
| P | 6527 | hCoV-19/Denmark/DCGC-7193/2020\|EPI_ISL_619192\|2020 | .................... | .................... | ..................... | .................... | .................... | .................... |
| P | 6528 | hCoV-19/Denmark/DCGC-7194/2020\|EPI_ISL_619193\|2020 | .................... | .................... | ..................... | .................... | .................... | .................... |
| P | 6529 | hCoV-19/Denmark/DCGC-7197/2020\|EPI_ISL_619194\|2020 | .................... | .................... | ..................... | .................... | .................... | .................... |
| P | 6530 | hCoV-19/Denmark/DCGC-7198/2020\|EPI_ISL_619195\|2020 | .................... | .................... | ..................... | .................... | .................... | .................... |
| P | 6531 | hCoV-19/Denmark/DCGC-7199/2020\|EPI_ISL_619196\|2020 | .................... | .................... | ..................... | .................... | .................... | .................... |
| P | 6532 | hCoV-19/Denmark/DCGC-7200/2020\|EPI_ISL_619197\|2020 | .................... | .................... | ..................... | .................... | .................... | .................... |
| P | 6533 | hCoV-19/Denmark/DCGC-7203/2020\|EPI_ISL_619198\|2020 | .................... | .................... | ..................... | .................... | .................... | .................... |
| P | 6534 | hCoV-19/Denmark/DCGC-7205/2020\|EPI_ISL_619199\|2020 | .................... | .................... | ..................... | .................... | .................... | .................... |
| P | 6535 | hCoV-19/Denmark/DCGC-6217/2020\|EPI_ISL_620120\|2020 | .................... | .................... | ..................... | .................... | .................... | .................... |
| P | 6536 | hCoV-19/Denmark/DCGC-6220/2020\|EPI_ISL_620123\|2020 | .................... | .................... | ..................... | .................... | .................... | .................... |
| P | 6537 | hCoV-19/Denmark/DCGC-6221/2020\|EPI_ISL_620124\|2020 | .................... | .................... | ..................... | .................... | .................... | .................... |
| P | 6538 | hCoV-19/Denmark/DCGC-6222/2020\|EPI_ISL_620125\|2020 | .................... | .................... | ..................... | .................... | .................... | .................... |
| P | 6539 | hCoV-19/Denmark/DCGC-6223/2020\|EPI_ISL_620126\|2020 | .................... | .................... | ..................... | .................... | .................... | .................... |
| P | 6540 | hCoV-19/Denmark/DCGC-1758/2020\|EPI_ISL_620127\|2020 | .................... | .................... | ..................... | .................... | .................... | .................... |
| P | 6541 | hCoV-19/Denmark/DCGC-1759/2020\|EPI_ISL_620128\|2020 | .................... | .................... | ..................... | .................... | .................... | .................... |

| | | | | | | | | |
|---|---|---|---|---|---|---|---|---|
| P | 6542 | hCoV-19/Denmark/DCGC-1706/2020\|EPI_ISL_620130\|2020 | .................... | .................... | ..................... | .................... | .................... | .................... |
| P | 6543 | hCoV-19/Denmark/DCGC-1747/2020\|EPI_ISL_620132\|2020 | .................... | .................... | ..................... | .................... | .................... | .................... |
| P | 6544 | hCoV-19/Denmark/DCGC-1749/2020\|EPI_ISL_620133\|2020 | .................... | .................... | ..................... | .................... | .................... | .................... |
| P | 6545 | hCoV-19/Denmark/DCGC-1750/2020\|EPI_ISL_620134\|2020 | .................... | .................... | ..................... | .................... | .................... | .................... |
| P | 6546 | hCoV-19/Denmark/DCGC-1753/2020\|EPI_ISL_620135\|2020 | .................... | .................... | ..................... | .................... | .................... | .................... |
| P | 6547 | hCoV-19/Denmark/DCGC-1756/2020\|EPI_ISL_620136\|2020 | .................... | .................... | ..................... | .................... | .................... | .................... |
| P | 6548 | hCoV-19/Denmark/DCGC-1751/2020\|EPI_ISL_620139\|2020 | .................... | .................... | ..................... | .................... | .................... | .................... |
| P | 6549 | hCoV-19/Denmark/DCGC-1754/2020\|EPI_ISL_620140\|2020 | .................... | .................... | ..................... | .................... | .................... | .................... |
| P | 6550 | hCoV-19/Denmark/DCGC-1757/2020\|EPI_ISL_620141\|2020 | .................... | .................... | ..................... | .................... | .................... | .................... |
| P | 6551 | hCoV-19/Denmark/DCGC-1760/2020\|EPI_ISL_620142\|2020 | .................... | .................... | ..................... | .................... | .................... | .................... |
| P | 6552 | hCoV-19/Denmark/DCGC-1282/2020\|EPI_ISL_620145\|2020 | .................... | .................... | ..................... | .................... | .................... | .................... |
| P | 6553 | hCoV-19/Denmark/DCGC-1284/2020\|EPI_ISL_620146\|2020 | .................... | .................... | ..................... | .................... | .................... | .................... |
| P | 6554 | hCoV-19/Denmark/DCGC-5883/2020\|EPI_ISL_620148\|2020 | .................... | .................... | ..................... | .................... | .................... | .................... |
| P | 6555 | hCoV-19/Denmark/DCGC-5884/2020\|EPI_ISL_620149\|2020 | .................... | .................... | ..................... | .................... | .................... | .................... |
| P | 6556 | hCoV-19/Denmark/DCGC-5885/2020\|EPI_ISL_620150\|2020 | .................... | .................... | ..................... | .................... | .................... | .................... |
| P | 6557 | hCoV-19/Denmark/DCGC-5886/2020\|EPI_ISL_620151\|2020 | .................... | .................... | ..................... | .................... | .................... | .................... |
| P | 6558 | hCoV-19/Denmark/DCGC-5888/2020\|EPI_ISL_620153\|2020 | .................... | .................... | ..................... | .................... | .................... | .................... |
| P | 6559 | hCoV-19/Denmark/DCGC-5889/2020\|EPI_ISL_620154\|2020 | .................... | .................... | ..................... | .................... | .................... | .................... |
| P | 6560 | hCoV-19/Denmark/DCGC-5890/2020\|EPI_ISL_620155\|2020 | .................... | .................... | ..................... | .................... | .................... | .................... |
| P | 6561 | hCoV-19/Denmark/DCGC-5891/2020\|EPI_ISL_620156\|2020 | .................... | .................... | ..................... | .................... | .................... | .................... |
| P | 6562 | hCoV-19/Denmark/DCGC-5892/2020\|EPI_ISL_620157\|2020 | .................... | .................... | ..................... | .................... | .................... | .................... |
| P | 6563 | hCoV-19/Denmark/DCGC-5893/2020\|EPI_ISL_620158\|2020 | .................... | .................... | ..................... | .................... | .................... | .................... |
| P | 6564 | hCoV-19/Denmark/DCGC-5894/2020\|EPI_ISL_620159\|2020 | .................... | .................... | ..................... | .................... | .................... | .................... |
| P | 6565 | hCoV-19/Denmark/DCGC-5895/2020\|EPI_ISL_620160\|2020 | .................... | .................... | ..................... | .................... | .................... | .................... |
| P | 6566 | hCoV-19/Denmark/DCGC-5896/2020\|EPI_ISL_620161\|2020 | .................... | .................... | ..................... | .................... | .................... | .................... |
| P | 6567 | hCoV-19/Denmark/DCGC-5897/2020\|EPI_ISL_620162\|2020 | .................... | .................... | ..................... | .................... | .................... | .................... |
| P | 6568 | hCoV-19/Denmark/DCGC-5898/2020\|EPI_ISL_620163\|2020 | .................... | .................... | ..................... | .................... | .................... | .................... |
| P | 6569 | hCoV-19/Denmark/DCGC-5902/2020\|EPI_ISL_620167\|2020 | .................... | .................... | ..................... | .................... | .................... | .................... |
| P | 6570 | hCoV-19/Denmark/DCGC-5904/2020\|EPI_ISL_620169\|2020 | .................... | .................... | ..................... | .................... | .................... | .................... |
| P | 6571 | hCoV-19/Denmark/DCGC-5905/2020\|EPI_ISL_620170\|2020 | .................... | .................... | ..................... | .................... | .................... | .................... |
| P | 6572 | hCoV-19/Denmark/DCGC-5906/2020\|EPI_ISL_620171\|2020 | .................... | .................... | ..................... | .................... | .................... | .................... |
| P | 6573 | hCoV-19/Denmark/DCGC-5909/2020\|EPI_ISL_620174\|2020 | .................... | .................... | ..................... | .................... | .................... | .................... |
| P | 6574 | hCoV-19/Denmark/DCGC-5911/2020\|EPI_ISL_620176\|2020 | .................... | .A.................. | ..................... | .................... | .................... | .................... |
| P | 6575 | hCoV-19/Denmark/DCGC-5912/2020\|EPI_ISL_620177\|2020 | .................... | .................... | ..................... | .................... | .................... | .................... |
| P | 6576 | hCoV-19/Denmark/DCGC-5913/2020\|EPI_ISL_620178\|2020 | .................... | .................... | ..................... | .................... | .................... | .................... |
| P | 6577 | hCoV-19/Denmark/DCGC-5914/2020\|EPI_ISL_620179\|2020 | .................... | .................... | ..................... | .................... | .................... | .................... |
| P | 6578 | hCoV-19/Denmark/DCGC-5915/2020\|EPI_ISL_620180\|2020 | .................... | .................... | ..................... | .................... | .................... | .................... |
| P | 6579 | hCoV-19/Denmark/DCGC-5918/2020\|EPI_ISL_620183\|2020 | .................... | .................... | ..................... | .................... | .................... | .................... |
| P | 6580 | hCoV-19/Denmark/DCGC-5919/2020\|EPI_ISL_620184\|2020 | .................... | .................... | ..................... | .................... | .................... | .................... |
| P | 6581 | hCoV-19/Denmark/DCGC-5920/2020\|EPI_ISL_620185\|2020 | .................... | .................... | ..................... | .................... | .................... | .................... |
| P | 6582 | hCoV-19/Denmark/DCGC-5921/2020\|EPI_ISL_620186\|2020 | .................... | .................... | ..................... | .................... | .................... | .................... |
| P | 6583 | hCoV-19/Denmark/DCGC-5922/2020\|EPI_ISL_620187\|2020 | .................... | .................... | ..................... | .................... | .................... | .................... |
| P | 6584 | hCoV-19/Denmark/DCGC-5924/2020\|EPI_ISL_620189\|2020 | .................... | .................... | ..................... | .................... | .................... | .................... |
| P | 6585 | hCoV-19/Denmark/DCGC-5927/2020\|EPI_ISL_620192\|2020 | .................... | .................... | ..................... | .................... | .................... | .................... |
| P | 6586 | hCoV-19/Denmark/DCGC-5928/2020\|EPI_ISL_620193\|2020 | .................... | .................... | ..................... | .................... | .................... | .................... |
| P | 6587 | hCoV-19/Denmark/DCGC-5929/2020\|EPI_ISL_620194\|2020 | .................... | .................... | ..................... | .................... | .................... | .................... |
| P | 6588 | hCoV-19/Denmark/DCGC-5930/2020\|EPI_ISL_620195\|2020 | .................... | .................... | ..................... | .................... | .................... | .................... |
| P | 6589 | hCoV-19/Denmark/DCGC-5931/2020\|EPI_ISL_620196\|2020 | .................... | .................... | ..................... | .................... | .................... | .................... |
| P | 6590 | hCoV-19/Denmark/DCGC-5932/2020\|EPI_ISL_620197\|2020 | .................... | .................... | ..................... | .................... | .................... | .................... |
| P | 6591 | hCoV-19/Denmark/DCGC-5933/2020\|EPI_ISL_620198\|2020 | .................... | .................... | ..................... | .................... | .................... | .................... |
| P | 6592 | hCoV-19/Denmark/DCGC-4909/2020\|EPI_ISL_621447\|2020 | .................... | .................... | ..................... | .................... | .................... | .................... |
| P | 6593 | hCoV-19/Denmark/DCGC-4910/2020\|EPI_ISL_621448\|2020 | .................... | .................... | ..................... | .................... | .................... | .................... |
| P | 6594 | hCoV-19/Denmark/DCGC-4911/2020\|EPI_ISL_621449\|2020 | .................... | .................... | ..................... | .................... | .................... | .................... |
| P | 6595 | hCoV-19/Denmark/DCGC-4912/2020\|EPI_ISL_621450\|2020 | .................... | .................... | ..................... | .................... | .................... | .................... |
| P | 6596 | hCoV-19/Denmark/DCGC-4913/2020\|EPI_ISL_621451\|2020 | .................... | .................... | ..................... | .................... | .................... | .................... |
| P | 6597 | hCoV-19/Denmark/DCGC-4914/2020\|EPI_ISL_621452\|2020 | .................... | .................... | ..................... | .................... | .................... | .................... |
| P | 6598 | hCoV-19/Denmark/DCGC-4915/2020\|EPI_ISL_621453\|2020 | .................... | .................... | ..................... | .................... | .................... | .................... |
| P | 6599 | hCoV-19/Denmark/DCGC-4917/2020\|EPI_ISL_621455\|2020 | .................... | .................... | ..................... | .................... | .................... | .................... |
| P | 6600 | hCoV-19/Denmark/DCGC-4918/2020\|EPI_ISL_621456\|2020 | .................... | .................... | ..................... | .................... | .................... | .................... |
| P | 6601 | hCoV-19/Denmark/DCGC-4919/2020\|EPI_ISL_621457\|2020 | .................... | .................... | ..................... | .................... | .................... | .................... |
| P | 6602 | hCoV-19/Denmark/DCGC-4920/2020\|EPI_ISL_621458\|2020 | .................... | .................... | ..................... | .................... | .................... | .................... |
| P | 6603 | hCoV-19/Denmark/DCGC-4921/2020\|EPI_ISL_621459\|2020 | .................... | .................... | ..................... | .................... | .................... | .................... |
| P | 6604 | hCoV-19/Denmark/DCGC-4922/2020\|EPI_ISL_621460\|2020 | .................... | .................... | ..................... | .................... | .................... | .................... |
| P | 6605 | hCoV-19/Denmark/DCGC-4923/2020\|EPI_ISL_621461\|2020 | .................... | .................... | ..................... | .................... | .................... | .................... |
| P | 6606 | hCoV-19/Denmark/DCGC-4924/2020\|EPI_ISL_621462\|2020 | .................... | .................... | ..................... | .................... | .................... | .................... |
| P | 6607 | hCoV-19/Denmark/DCGC-4925/2020\|EPI_ISL_621463\|2020 | .................... | .A.................. | ..................... | .................... | .................... | .................... |
| P | 6608 | hCoV-19/Denmark/DCGC-4926/2020\|EPI_ISL_621464\|2020 | .................... | .................... | ..................... | .................... | .................... | .................... |
| P | 6609 | hCoV-19/Denmark/DCGC-4927/2020\|EPI_ISL_621465\|2020 | .................... | .................... | ..................... | .................... | .................... | .................... |
| P | 6610 | hCoV-19/Denmark/DCGC-4928/2020\|EPI_ISL_621466\|2020 | .................... | .................... | ..................... | .................... | .................... | .................... |
| P | 6611 | hCoV-19/Denmark/DCGC-4930/2020\|EPI_ISL_621468\|2020 | .................... | .................... | ..................... | .................... | ......T............. | .................... |
| P | 6612 | hCoV-19/Denmark/DCGC-4931/2020\|EPI_ISL_621469\|2020 | .................... | .................... | ..................... | .................... | .................... | .................... |
| P | 6613 | hCoV-19/Denmark/DCGC-4932/2020\|EPI_ISL_621470\|2020 | .................... | .................... | ..................... | .................... | .................... | .................... |
| P | 6614 | hCoV-19/Denmark/DCGC-4933/2020\|EPI_ISL_621471\|2020 | .................... | .................... | ..................... | .................... | .................... | .................... |
| P | 6615 | hCoV-19/Denmark/DCGC-4934/2020\|EPI_ISL_621472\|2020 | .................... | .................... | ..................... | .................... | .................... | .................... |
| P | 6616 | hCoV-19/Denmark/DCGC-4935/2020\|EPI_ISL_621473\|2020 | .................... | .................... | ..................... | .................... | .................... | .................... |
| P | 6617 | hCoV-19/Denmark/DCGC-4936/2020\|EPI_ISL_621474\|2020 | .................... | .................... | ..................... | .................... | .................... | .................... |
| P | 6618 | hCoV-19/Denmark/DCGC-4937/2020\|EPI_ISL_621475\|2020 | .................... | .................... | ..................... | .................... | .................... | .................... |
| P | 6619 | hCoV-19/Denmark/DCGC-4938/2020\|EPI_ISL_621476\|2020 | .................... | .................... | ..................... | .................... | .................... | .................... |
| P | 6620 | hCoV-19/Denmark/DCGC-4939/2020\|EPI_ISL_621477\|2020 | .................... | .................... | ..................... | .................... | .................... | .................... |
| P | 6621 | hCoV-19/Denmark/DCGC-4940/2020\|EPI_ISL_621478\|2020 | .................... | .................... | ..................... | .................... | .................... | .................... |

| | | | | | | | | |
|---|---|---|---|---|---|---|---|---|
| P | 6622 | hCoV-19/Denmark/DCGC-4941/2020\|EPI_ISL_621479\|2020 | ···················· | ···················· | ···················· | ···················· | ···················· | ···················· |
| P | 6623 | hCoV-19/Denmark/DCGC-4942/2020\|EPI_ISL_621480\|2020 | ···················· | ···················· | ···················· | ···················· | ···················· | ···················· |
| P | 6624 | hCoV-19/Denmark/DCGC-4943/2020\|EPI_ISL_621481\|2020 | ···················· | ···················· | ···················· | ···················· | ···················· | ···················· |
| P | 6625 | hCoV-19/Denmark/DCGC-4944/2020\|EPI_ISL_621482\|2020 | ···················· | ···················· | ···················· | ···················· | ···················· | ···················· |
| P | 6626 | hCoV-19/Denmark/DCGC-4945/2020\|EPI_ISL_621483\|2020 | ···················· | ···················· | ···················· | ···················· | ···················· | ···················· |
| P | 6627 | hCoV-19/Denmark/DCGC-4946/2020\|EPI_ISL_621484\|2020 | ···················· | ···················· | ···················· | ···················· | ···················· | ···················· |
| P | 6628 | hCoV-19/Denmark/DCGC-4947/2020\|EPI_ISL_621485\|2020 | ···················· | ···················· | ···················· | ···················· | ···················· | ···················· |
| P | 6629 | hCoV-19/Denmark/DCGC-4948/2020\|EPI_ISL_621486\|2020 | ···················· | ···················· | ···················· | ···················· | ···················· | ···················· |
| P | 6630 | hCoV-19/Denmark/DCGC-4949/2020\|EPI_ISL_621487\|2020 | ···················· | ···················· | ···················· | ···················· | ···················· | ···················· |
| P | 6631 | hCoV-19/Denmark/DCGC-4951/2020\|EPI_ISL_621489\|2020 | ···················· | ···················· | ···················· | ···················· | ···················· | ···················· |
| P | 6632 | hCoV-19/Denmark/DCGC-4952/2020\|EPI_ISL_621490\|2020 | ···················· | ···················· | ···················· | ···················· | ···················· | ···················· |
| P | 6633 | hCoV-19/Denmark/DCGC-4953/2020\|EPI_ISL_621491\|2020 | ···················· | ···················· | ···················· | ···················· | ···················· | ···················· |
| P | 6634 | hCoV-19/Denmark/DCGC-4954/2020\|EPI_ISL_621492\|2020 | ···················· | ···················· | ···················· | ···················· | ···················· | ···················· |
| P | 6635 | hCoV-19/Denmark/DCGC-4955/2020\|EPI_ISL_621493\|2020 | ···················· | ···················· | ···················· | ···················· | ···················· | ···················· |
| P | 6636 | hCoV-19/Denmark/DCGC-4956/2020\|EPI_ISL_621494\|2020 | ···················· | ···················· | ···················· | ···················· | ···················· | ···················· |
| P | 6637 | hCoV-19/Denmark/DCGC-4957/2020\|EPI_ISL_621495\|2020 | ···················· | ···················· | ···················· | ···················· | ···················· | ···················· |
| P | 6638 | hCoV-19/Denmark/DCGC-4958/2020\|EPI_ISL_621496\|2020 | ···················· | ···················· | ···················· | ···················· | ···················· | ···················· |
| P | 6639 | hCoV-19/Denmark/DCGC-4959/2020\|EPI_ISL_621497\|2020 | ···················· | ···················· | ···················· | ···················· | ···················· | ···················· |
| P | 6640 | hCoV-19/Denmark/DCGC-4960/2020\|EPI_ISL_621498\|2020 | ···················· | ···················· | ···················· | ···················· | ···················· | ···················· |
| P | 6641 | hCoV-19/Denmark/DCGC-4961/2020\|EPI_ISL_621499\|2020 | ···················· | ···················· | ···················· | ···················· | ···················· | ···················· |
| P | 6642 | hCoV-19/New Zealand/20CV0510/2020\|EPI_ISL_622787\|2 | ···················· | ···················· | ···················· | ···················· | ···················· | ···················· |
| P | 6643 | hCoV-19/New Zealand/20CV0506/2020\|EPI_ISL_622792\|2 | ···················· | ·A·················· | ···················· | ···················· | ···················· | ··A················· |
| P | 6644 | hCoV-19/New Zealand/20CV0297/2020\|EPI_ISL_622794\|2 | ···················· | ···················· | ···················· | ···················· | ···················· | ···················· |
| P | 6645 | hCoV-19/New Zealand/20CV0298/2020\|EPI_ISL_622795\|2 | ···················· | ···················· | ···················· | ···················· | ···················· | ···················· |
| P | 6646 | hCoV-19/New Zealand/20CV0299/2020\|EPI_ISL_622796\|2 | ···················· | ···················· | ···················· | ···················· | ···················· | ···················· |
| P | 6647 | hCoV-19/New Zealand/20CV0302/2020\|EPI_ISL_622797\|2 | ···················· | ···················· | ···················· | ···················· | ···················· | ···················· |
| P | 6648 | hCoV-19/New Zealand/20CV0303/2020\|EPI_ISL_622798\|2 | ···················· | ···················· | ···················· | ···················· | ···················· | ···················· |
| P | 6649 | hCoV-19/New Zealand/20CV0306/2020\|EPI_ISL_622799\|2 | ···················· | ···················· | ···················· | ···················· | ···················· | ···················· |
| P | 6650 | hCoV-19/Denmark/DCGC-9013/2020\|EPI_ISL_626210\|2020 | ···················· | ···················· | ···················· | ···················· | ···················· | ···················· |
| P | 6651 | hCoV-19/Germany/NW-HHU-212/2020\|EPI_ISL_626211\|202 | ···················· | ···················· | ···················· | ···················· | ···················· | ···················· |
| P | 6652 | hCoV-19/Germany/NW-HHU-213/2020\|EPI_ISL_626212\|202 | ···················· | ···················· | ···················· | ···················· | ···················· | ···················· |
| P | 6653 | hCoV-19/Germany/NW-HHU-215/2020\|EPI_ISL_626214\|202 | ···················· | ···················· | ···················· | ···················· | ···················· | ···················· |
| P | 6654 | hCoV-19/Germany/NW-HHU-217/2020\|EPI_ISL_626216\|202 | ···················· | ···················· | ···················· | ···················· | ···················· | ···················· |
| P | 6655 | hCoV-19/Germany/NW-HHU-218/2020\|EPI_ISL_626217\|202 | ···················· | ···················· | ···················· | ···················· | ···················· | ···················· |
| P | 6656 | hCoV-19/Germany/NW-HHU-220/2020\|EPI_ISL_626219\|202 | ···················· | ···················· | ···················· | ···················· | ···················· | ···················· |
| P | 6657 | hCoV-19/USA/NY-PHL-000011/2020\|EPI_ISL_614294\|2020 | ···················· | ···················· | ···················· | ···················· | ···················· | ···················· |
| P | 6658 | hCoV-19/Faroe Islands/HFS-119/2020\|EPI_ISL_614298\| | ···················· | ···················· | ···················· | ···················· | ···················· | ···················· |
| P | 6659 | hCoV-19/Denmark/DCGC-3722/2020\|EPI_ISL_615501\|2020 | ···················· | ···················· | ···················· | ···················· | ···················· | ···················· |
| P | 6660 | hCoV-19/Denmark/DCGC-3723/2020\|EPI_ISL_615502\|2020 | ···················· | ···················· | ···················· | ···················· | ···················· | ···················· |
| P | 6661 | hCoV-19/Denmark/DCGC-6310/2020\|EPI_ISL_615503\|2020 | ···················· | ···················· | ···················· | ···················· | ···················· | ···················· |
| P | 6662 | hCoV-19/Denmark/DCGC-3724/2020\|EPI_ISL_615504\|2020 | ···················· | ···················· | ···················· | ···················· | ···················· | ···················· |
| P | 6663 | hCoV-19/Denmark/DCGC-3725/2020\|EPI_ISL_615505\|2020 | ···················· | ···················· | ···················· | ···················· | ···················· | ···················· |
| P | 6664 | hCoV-19/Denmark/DCGC-3727/2020\|EPI_ISL_615507\|2020 | ···················· | ···················· | ···················· | ···················· | ···················· | ···················· |
| P | 6665 | hCoV-19/Denmark/DCGC-3728/2020\|EPI_ISL_615508\|2020 | ···················· | ···················· | ···················· | ···················· | ···················· | ···················· |
| P | 6666 | hCoV-19/Denmark/DCGC-3729/2020\|EPI_ISL_615509\|2020 | ···················· | ···················· | ···················· | ···················· | ···················· | ···················· |
| P | 6667 | hCoV-19/Denmark/DCGC-3730/2020\|EPI_ISL_615510\|2020 | ···················· | ···················· | ···················· | ···················· | ···················· | ···················· |
| P | 6668 | hCoV-19/Denmark/DCGC-3731/2020\|EPI_ISL_615511\|2020 | ···················· | ···················· | ···················· | ···················· | ···················· | ···················· |
| P | 6669 | hCoV-19/Denmark/DCGC-3732/2020\|EPI_ISL_615512\|2020 | ···················· | ···················· | ···················· | ···················· | ···················· | ···················· |
| P | 6670 | hCoV-19/Denmark/DCGC-3733/2020\|EPI_ISL_615513\|2020 | ···················· | ···················· | ···················· | ···················· | ···················· | ···················· |
| P | 6671 | hCoV-19/Denmark/DCGC-6311/2020\|EPI_ISL_615514\|2020 | ···················· | ···················· | ···················· | ···················· | ···················· | ···················· |
| P | 6672 | hCoV-19/Denmark/DCGC-6312/2020\|EPI_ISL_615515\|2020 | ···················· | ···················· | ···················· | ···················· | ···················· | ···················· |
| P | 6673 | hCoV-19/Denmark/DCGC-4316/2020\|EPI_ISL_615516\|2020 | ···················· | ···················· | ···················· | ···················· | ···················· | ···················· |
| P | 6674 | hCoV-19/Denmark/DCGC-4317/2020\|EPI_ISL_615517\|2020 | ···················· | ···················· | ···················· | ···················· | ···················· | ···················· |
| P | 6675 | hCoV-19/Denmark/DCGC-4318/2020\|EPI_ISL_615518\|2020 | ···················· | ···················· | ···················· | ···················· | ···················· | ···················· |
| P | 6676 | hCoV-19/Denmark/DCGC-4319/2020\|EPI_ISL_615519\|2020 | ···················· | ···················· | ···················· | ···················· | ···················· | ···················· |
| P | 6677 | hCoV-19/Denmark/DCGC-4320/2020\|EPI_ISL_615520\|2020 | ···················· | ···················· | ···················· | ···················· | ···················· | ···················· |
| P | 6678 | hCoV-19/Denmark/DCGC-4321/2020\|EPI_ISL_615521\|2020 | ···················· | ···················· | ···················· | ···················· | ···················· | ···················· |
| P | 6679 | hCoV-19/Denmark/DCGC-4322/2020\|EPI_ISL_615522\|2020 | ···················· | ···················· | ···················· | ···················· | ···················· | ···················· |
| P | 6680 | hCoV-19/Denmark/DCGC-4323/2020\|EPI_ISL_615523\|2020 | ···················· | ···················· | ···················· | ···················· | ···················· | ···················· |
| P | 6681 | hCoV-19/Denmark/DCGC-4324/2020\|EPI_ISL_615524\|2020 | ···················· | ···················· | ···················· | ···················· | ···················· | ···················· |
| P | 6682 | hCoV-19/Denmark/DCGC-4325/2020\|EPI_ISL_615525\|2020 | ···················· | ···················· | ···················· | ···················· | ···················· | ···················· |
| P | 6683 | hCoV-19/Denmark/DCGC-4326/2020\|EPI_ISL_615526\|2020 | ···················· | ···················· | ···················· | ···················· | ···················· | ···················· |
| P | 6684 | hCoV-19/Denmark/DCGC-4327/2020\|EPI_ISL_615527\|2020 | ···················· | ···················· | ···················· | ···················· | ···················· | ···················· |
| P | 6685 | hCoV-19/Denmark/DCGC-4328/2020\|EPI_ISL_615528\|2020 | ···················· | ···················· | ···················· | ···················· | ···················· | ···················· |
| P | 6686 | hCoV-19/Denmark/DCGC-4359/2020\|EPI_ISL_615561\|2020 | ···················· | ···················· | ···················· | ···················· | ···················· | ···················· |
| P | 6687 | hCoV-19/Denmark/DCGC-4360/2020\|EPI_ISL_615562\|2020 | ···················· | ···················· | ···················· | ···················· | ···················· | ···················· |
| P | 6688 | hCoV-19/Denmark/DCGC-4361/2020\|EPI_ISL_615563\|2020 | ···················· | ·A·················· | ···················· | ···················· | ···················· | ···················· |
| P | 6689 | hCoV-19/Denmark/DCGC-6315/2020\|EPI_ISL_615564\|2020 | ···················· | ···················· | ···················· | ···················· | ···················· | ···················· |
| P | 6690 | hCoV-19/Denmark/DCGC-4368/2020\|EPI_ISL_615571\|2020 | ···················· | ···················· | ···················· | ···················· | ·····T·············· | ···················· |
| P | 6691 | hCoV-19/Denmark/DCGC-4369/2020\|EPI_ISL_615572\|2020 | ···················· | ···················· | ···················· | ···················· | ···················· | ···················· |
| P | 6692 | hCoV-19/Denmark/DCGC-4371/2020\|EPI_ISL_615574\|2020 | ···················· | ···················· | ···················· | ···················· | ···················· | ···················· |
| P | 6693 | hCoV-19/Denmark/DCGC-4372/2020\|EPI_ISL_615575\|2020 | ···················· | ···················· | ···················· | ···················· | ···················· | ···················· |
| P | 6694 | hCoV-19/Denmark/DCGC-4373/2020\|EPI_ISL_615576\|2020 | ···················· | ···················· | ···················· | ···················· | ···················· | ···················· |
| P | 6695 | hCoV-19/Denmark/DCGC-1718/2020\|EPI_ISL_615577\|2020 | ···················· | ···················· | ···················· | ···················· | ···················· | ···················· |
| P | 6696 | hCoV-19/Denmark/DCGC-4374/2020\|EPI_ISL_615578\|2020 | ···················· | ···················· | ···················· | ···················· | ···················· | ···················· |
| P | 6697 | hCoV-19/Denmark/DCGC-4375/2020\|EPI_ISL_615579\|2020 | ···················· | ···················· | ···················· | ···················· | ···················· | ···················· |
| P | 6698 | hCoV-19/Denmark/DCGC-4376/2020\|EPI_ISL_615580\|2020 | ···················· | ···················· | ···················· | ···················· | ···················· | ···················· |
| P | 6699 | hCoV-19/Denmark/DCGC-4377/2020\|EPI_ISL_615581\|2020 | ···················· | ···················· | ···················· | ···················· | ···················· | ···················· |
| P | 6700 | hCoV-19/Denmark/DCGC-6316/2020\|EPI_ISL_615582\|2020 | ···················· | ···················· | ···················· | ···················· | ···················· | ···················· |
| P | 6701 | hCoV-19/Denmark/DCGC-3865/2020\|EPI_ISL_615583\|2020 | ···················· | ···················· | ···················· | ···················· | ···················· | ···················· |

| | | | | | | | | |
|---|---|---|---|---|---|---|---|---|
| P | 6702 | hCoV-19/Denmark/DCGC-3866/2020\|EPI_ISL_615584\|2020 | ···················· | ···················· | ····················· | ···················· | ···················· | ···················· |
| P | 6703 | hCoV-19/Denmark/DCGC-3868/2020\|EPI_ISL_615586\|2020 | ···················· | ···················· | ····················· | ···················· | ···················· | ···················· |
| P | 6704 | hCoV-19/Denmark/DCGC-3869/2020\|EPI_ISL_615587\|2020 | ···················· | ···················· | ····················· | ···················· | ···················· | ···················· |
| P | 6705 | hCoV-19/Denmark/DCGC-3870/2020\|EPI_ISL_615588\|2020 | ···················· | ···················· | ····················· | ···················· | ···················· | ···················· |
| P | 6706 | hCoV-19/Denmark/DCGC-3871/2020\|EPI_ISL_615589\|2020 | ···················· | ···················· | ····················· | ···················· | ···················· | ···················· |
| P | 6707 | hCoV-19/Denmark/DCGC-3872/2020\|EPI_ISL_615590\|2020 | ···················· | ···················· | ····················· | ···················· | ···················· | ···················· |
| P | 6708 | hCoV-19/Denmark/DCGC-3873/2020\|EPI_ISL_615591\|2020 | ···················· | ···················· | ····················· | ···················· | ···················· | ···················· |
| P | 6709 | hCoV-19/Denmark/DCGC-3874/2020\|EPI_ISL_615592\|2020 | ···················· | ···················· | ····················· | ···················· | ···················· | ···················· |
| P | 6710 | hCoV-19/Denmark/DCGC-3876/2020\|EPI_ISL_615594\|2020 | ···················· | ···················· | ····················· | ···················· | ···················· | ···················· |
| P | 6711 | hCoV-19/Denmark/DCGC-3877/2020\|EPI_ISL_615595\|2020 | ···················· | ···················· | ····················· | ···················· | ···················· | ···················· |
| P | 6712 | hCoV-19/Denmark/DCGC-3878/2020\|EPI_ISL_615596\|2020 | ···················· | ···················· | ····················· | ···················· | ···················· | ···················· |
| P | 6713 | hCoV-19/Denmark/DCGC-3879/2020\|EPI_ISL_615597\|2020 | ···················· | ···················· | ····················· | ···················· | ···················· | ···················· |
| P | 6714 | hCoV-19/Denmark/DCGC-3880/2020\|EPI_ISL_615598\|2020 | ···················· | ···················· | ····················· | ···················· | ···················· | ···················· |
| P | 6715 | hCoV-19/Denmark/DCGC-3881/2020\|EPI_ISL_615599\|2020 | ···················· | ···················· | ····················· | ···················· | ···················· | ···················· |
| P | 6716 | hCoV-19/Denmark/DCGC-3038/2020\|EPI_ISL_616816\|2020 | ···················· | ···················· | ····················· | ···················· | ···················· | ···················· |
| P | 6717 | hCoV-19/Denmark/DCGC-3039/2020\|EPI_ISL_616817\|2020 | ···················· | ···················· | ····················· | ···················· | ···················· | ···················· |
| P | 6718 | hCoV-19/Denmark/DCGC-3040/2020\|EPI_ISL_616818\|2020 | ···················· | ···················· | ····················· | ···················· | ···················· | ···················· |
| P | 6719 | hCoV-19/Denmark/DCGC-3042/2020\|EPI_ISL_616820\|2020 | ···················· | ···················· | ····················· | ···················· | ···················· | ···················· |
| P | 6720 | hCoV-19/Denmark/DCGC-3043/2020\|EPI_ISL_616821\|2020 | ···················· | ···················· | ····················· | ···················· | ···················· | ···················· |
| P | 6721 | hCoV-19/Denmark/DCGC-3242/2020\|EPI_ISL_616822\|2020 | ···················· | ···················· | ····················· | ···················· | ···················· | ···················· |
| P | 6722 | hCoV-19/Denmark/DCGC-3243/2020\|EPI_ISL_616823\|2020 | ···················· | ···················· | ····················· | ···················· | ···················· | ···················· |
| P | 6723 | hCoV-19/Denmark/DCGC-3044/2020\|EPI_ISL_616824\|2020 | ···················· | ···················· | ····················· | ···················· | ···················· | ···················· |
| P | 6724 | hCoV-19/Denmark/DCGC-3045/2020\|EPI_ISL_616825\|2020 | ···················· | ···················· | ····················· | ···················· | ···················· | ···················· |
| P | 6725 | hCoV-19/Denmark/DCGC-3047/2020\|EPI_ISL_616827\|2020 | ···················· | ···················· | ····················· | ···················· | ···················· | ···················· |
| P | 6726 | hCoV-19/Denmark/DCGC-3049/2020\|EPI_ISL_616829\|2020 | ···················· | ·A·················· | ····················· | ···················· | ···················· | ···················· |
| P | 6727 | hCoV-19/Denmark/DCGC-3050/2020\|EPI_ISL_616830\|2020 | ···················· | ···················· | ····················· | ···················· | ···················· | ···················· |
| P | 6728 | hCoV-19/Denmark/DCGC-3051/2020\|EPI_ISL_616831\|2020 | ···················· | ···················· | ····················· | ···················· | ···················· | ···················· |
| P | 6729 | hCoV-19/Denmark/DCGC-3052/2020\|EPI_ISL_616832\|2020 | ···················· | ···················· | ····················· | ···················· | ···················· | ···················· |
| P | 6730 | hCoV-19/Denmark/DCGC-3053/2020\|EPI_ISL_616833\|2020 | ···················· | ···················· | ····················· | ···················· | ···················· | ···················· |
| P | 6731 | hCoV-19/Denmark/DCGC-3054/2020\|EPI_ISL_616834\|2020 | ···················· | ···················· | ····················· | ···················· | ···················· | ···················· |
| P | 6732 | hCoV-19/Denmark/DCGC-3055/2020\|EPI_ISL_616835\|2020 | ···················· | ···················· | ····················· | ···················· | ···················· | ···················· |
| P | 6733 | hCoV-19/Denmark/DCGC-3056/2020\|EPI_ISL_616836\|2020 | ···················· | ·A·················· | ····················· | ···················· | ···················· | ··A················· |
| P | 6734 | hCoV-19/Denmark/DCGC-3057/2020\|EPI_ISL_616837\|2020 | ···················· | ···················· | ····················· | ···················· | ···················· | ···················· |
| P | 6735 | hCoV-19/Denmark/DCGC-3058/2020\|EPI_ISL_616838\|2020 | ···················· | ···················· | ····················· | ···················· | ···················· | ···················· |
| P | 6736 | hCoV-19/Denmark/DCGC-3059/2020\|EPI_ISL_616839\|2020 | ···················· | ···················· | ····················· | ···················· | ···················· | ···················· |
| P | 6737 | hCoV-19/Denmark/DCGC-3060/2020\|EPI_ISL_616840\|2020 | ···················· | ···················· | ····················· | ···················· | ···················· | ···················· |
| P | 6738 | hCoV-19/Denmark/DCGC-3061/2020\|EPI_ISL_616841\|2020 | ···················· | ···················· | ····················· | ···················· | ···················· | ···················· |
| P | 6739 | hCoV-19/Denmark/DCGC-3062/2020\|EPI_ISL_616842\|2020 | ···················· | ···················· | ····················· | ···················· | ···················· | ···················· |
| P | 6740 | hCoV-19/Denmark/DCGC-3063/2020\|EPI_ISL_616843\|2020 | ···················· | ···················· | ····················· | ···················· | ···················· | ···················· |
| P | 6741 | hCoV-19/Denmark/DCGC-3064/2020\|EPI_ISL_616844\|2020 | ···················· | ···················· | ····················· | ···················· | ···················· | ···················· |
| P | 6742 | hCoV-19/Denmark/DCGC-3065/2020\|EPI_ISL_616845\|2020 | ···················· | ···················· | ····················· | ···················· | ···················· | ···················· |
| P | 6743 | hCoV-19/Denmark/DCGC-3066/2020\|EPI_ISL_616846\|2020 | ···················· | ···················· | ····················· | ···················· | ···················· | ···················· |
| P | 6744 | hCoV-19/Denmark/DCGC-3067/2020\|EPI_ISL_616847\|2020 | ···················· | ···················· | ····················· | ···················· | ···················· | ···················· |
| P | 6745 | hCoV-19/Denmark/DCGC-3068/2020\|EPI_ISL_616848\|2020 | ···················· | ···················· | ····················· | ···················· | ···················· | ···················· |
| P | 6746 | hCoV-19/Denmark/DCGC-3069/2020\|EPI_ISL_616849\|2020 | ···················· | ···················· | ····················· | ···················· | ···················· | ···················· |
| P | 6747 | hCoV-19/Denmark/DCGC-3070/2020\|EPI_ISL_616850\|2020 | ···················· | ···················· | ····················· | ···················· | ···················· | ···················· |
| P | 6748 | hCoV-19/Denmark/DCGC-3071/2020\|EPI_ISL_616851\|2020 | ···················· | ·A·················· | ····················· | ···················· | ···················· | ···················· |
| P | 6749 | hCoV-19/Denmark/DCGC-3072/2020\|EPI_ISL_616852\|2020 | ···················· | ···················· | ····················· | ···················· | ···················· | ···················· |
| P | 6750 | hCoV-19/Denmark/DCGC-3073/2020\|EPI_ISL_616853\|2020 | ···················· | ···················· | ····················· | ···················· | ···················· | ···················· |
| P | 6751 | hCoV-19/Denmark/DCGC-3074/2020\|EPI_ISL_616854\|2020 | ···················· | ···················· | ····················· | ···················· | ···················· | ···················· |
| P | 6752 | hCoV-19/Denmark/DCGC-3075/2020\|EPI_ISL_616855\|2020 | ···················· | ···················· | ····················· | ···················· | ···················· | ···················· |
| P | 6753 | hCoV-19/Denmark/DCGC-3076/2020\|EPI_ISL_616856\|2020 | ···················· | ···················· | ····················· | ···················· | ···················· | ···················· |
| P | 6754 | hCoV-19/Denmark/DCGC-3077/2020\|EPI_ISL_616857\|2020 | ···················· | ···················· | ····················· | ···················· | ···················· | ···················· |
| P | 6755 | hCoV-19/Denmark/DCGC-3078/2020\|EPI_ISL_616858\|2020 | ···················· | ···················· | ····················· | ···················· | ···················· | ···················· |
| P | 6756 | hCoV-19/Denmark/DCGC-3230/2020\|EPI_ISL_616890\|2020 | ···················· | ·A·················· | ····················· | ···················· | ···················· | ··A················· |
| P | 6757 | hCoV-19/Denmark/DCGC-3231/2020\|EPI_ISL_616891\|2020 | ···················· | ·A·················· | ····················· | ···················· | ···················· | ··A················· |
| P | 6758 | hCoV-19/Denmark/DCGC-3233/2020\|EPI_ISL_616893\|2020 | ···················· | ···················· | ····················· | ···················· | ···················· | ···················· |
| P | 6759 | hCoV-19/Denmark/DCGC-3234/2020\|EPI_ISL_616894\|2020 | ···················· | ·A·················· | ····················· | ···················· | ···················· | ···················· |
| P | 6760 | hCoV-19/Denmark/DCGC-3235/2020\|EPI_ISL_616895\|2020 | ···················· | ···················· | ····················· | ···················· | ···················· | ···················· |
| P | 6761 | hCoV-19/Denmark/DCGC-7345/2020\|EPI_ISL_619090\|2020 | ···················· | ···················· | ····················· | ···················· | ···················· | ···················· |
| P | 6762 | hCoV-19/Denmark/DCGC-7347/2020\|EPI_ISL_619091\|2020 | ···················· | ···················· | ····················· | ···················· | ···················· | ···················· |
| P | 6763 | hCoV-19/Denmark/DCGC-6178/2020\|EPI_ISL_620081\|2020 | ···················· | ···················· | ····················· | ···················· | ···················· | ···················· |
| P | 6764 | hCoV-19/Denmark/DCGC-6179/2020\|EPI_ISL_620082\|2020 | ···················· | ···················· | ····················· | ···················· | ···················· | ···················· |
| P | 6765 | hCoV-19/Denmark/DCGC-6180/2020\|EPI_ISL_620083\|2020 | ···················· | ···················· | ····················· | ···················· | ···················· | ···················· |
| P | 6766 | hCoV-19/Denmark/DCGC-6181/2020\|EPI_ISL_620084\|2020 | ···················· | ···················· | ····················· | ···················· | ···················· | ···················· |
| P | 6767 | hCoV-19/Denmark/DCGC-6182/2020\|EPI_ISL_620085\|2020 | ···················· | ···················· | ····················· | ···················· | ···················· | ···················· |
| P | 6768 | hCoV-19/Denmark/DCGC-6183/2020\|EPI_ISL_620086\|2020 | ···················· | ···················· | ····················· | ···················· | ···················· | ···················· |
| P | 6769 | hCoV-19/Denmark/DCGC-6184/2020\|EPI_ISL_620087\|2020 | ···················· | ···················· | ····················· | ···················· | ···················· | ···················· |
| P | 6770 | hCoV-19/Denmark/DCGC-6185/2020\|EPI_ISL_620088\|2020 | ···················· | ···················· | ····················· | ···················· | ···················· | ···················· |
| P | 6771 | hCoV-19/Denmark/DCGC-6186/2020\|EPI_ISL_620089\|2020 | ···················· | ·A·················· | ····················· | ···················· | ···················· | ···················· |
| P | 6772 | hCoV-19/Denmark/DCGC-6187/2020\|EPI_ISL_620090\|2020 | ···················· | ···················· | ····················· | ···················· | ···················· | ···················· |
| P | 6773 | hCoV-19/Denmark/DCGC-6188/2020\|EPI_ISL_620091\|2020 | ···················· | ···················· | ····················· | ···················· | ···················· | ···················· |
| P | 6774 | hCoV-19/Denmark/DCGC-6190/2020\|EPI_ISL_620093\|2020 | ···················· | ···················· | ····················· | ···················· | ···················· | ···················· |
| P | 6775 | hCoV-19/Denmark/DCGC-6191/2020\|EPI_ISL_620094\|2020 | ···················· | ···················· | ····················· | ···················· | ···················· | ···················· |
| P | 6776 | hCoV-19/Denmark/DCGC-6194/2020\|EPI_ISL_620097\|2020 | ···················· | ···················· | ····················· | ···················· | ···················· | ···················· |
| P | 6777 | hCoV-19/Denmark/DCGC-6195/2020\|EPI_ISL_620098\|2020 | ···················· | ···················· | ····················· | ···················· | ···················· | ···················· |
| P | 6778 | hCoV-19/Denmark/DCGC-4631/2020\|EPI_ISL_621300\|2020 | ···················· | ···················· | ····················· | ···················· | ···················· | ···················· |
| P | 6779 | hCoV-19/Denmark/DCGC-4632/2020\|EPI_ISL_621301\|2020 | ···················· | ···················· | ····················· | ···················· | ···················· | ···················· |
| P | 6780 | hCoV-19/Denmark/DCGC-4633/2020\|EPI_ISL_621302\|2020 | ···················· | ···················· | ····················· | ···················· | ···················· | ···················· |
| P | 6781 | hCoV-19/Denmark/DCGC-4634/2020\|EPI_ISL_621303\|2020 | ···················· | ···················· | ····················· | ···················· | ···················· | ···················· |

| | | | | | | | | |
|---|---|---|---|---|---|---|---|---|
| P | 6782 | hCoV-19/Denmark/DCGC-4519/2020\|EPI_ISL_621315\|2020 | ···················· | ···················· | ···················· | ···················· | ···················· | ···················· |
| P | 6783 | hCoV-19/Denmark/DCGC-4520/2020\|EPI_ISL_621316\|2020 | ···················· | ···················· | ···················· | ···················· | ···················· | ···················· |
| P | 6784 | hCoV-19/Denmark/DCGC-4521/2020\|EPI_ISL_621317\|2020 | ···················· | ···················· | ···················· | ···················· | ···················· | ···················· |
| P | 6785 | hCoV-19/Denmark/DCGC-4522/2020\|EPI_ISL_621318\|2020 | ···················· | ···················· | ···················· | ···················· | ···················· | ···················· |
| P | 6786 | hCoV-19/Denmark/DCGC-4523/2020\|EPI_ISL_621319\|2020 | ···················· | ···················· | ···················· | ···················· | ···················· | ···················· |
| P | 6787 | hCoV-19/Denmark/DCGC-1893/2020\|EPI_ISL_622613\|2020 | ···················· | ···················· | ···················· | ···················· | ···················· | ···················· |
| P | 6788 | hCoV-19/Denmark/DCGC-1899/2020\|EPI_ISL_622614\|2020 | ···················· | ···················· | ···················· | ···················· | ···················· | ···················· |
| P | 6789 | hCoV-19/Denmark/DCGC-3734/2020\|EPI_ISL_622615\|2020 | ···················· | ···················· | ···················· | ···················· | ···················· | ···················· |
| P | 6790 | hCoV-19/Denmark/DCGC-3739/2020\|EPI_ISL_622616\|2020 | ···················· | ···················· | ···················· | ···················· | ···················· | ···················· |
| P | 6791 | hCoV-19/Denmark/DCGC-3746/2020\|EPI_ISL_622617\|2020 | ···················· | ···················· | ···················· | ···················· | ···················· | ···················· |
| P | 6792 | hCoV-19/Denmark/DCGC-3752/2020\|EPI_ISL_622618\|2020 | ···················· | ···················· | ···················· | ···················· | ···················· | ···················· |
| P | 6793 | hCoV-19/Denmark/DCGC-3757/2020\|EPI_ISL_622619\|2020 | ···················· | ···················· | ···················· | ···················· | ···················· | ···················· |
| P | 6794 | hCoV-19/Denmark/DCGC-3760/2020\|EPI_ISL_622620\|2020 | ···················· | ···················· | ···················· | ···················· | ···················· | ···················· |
| P | 6795 | hCoV-19/Denmark/DCGC-3767/2020\|EPI_ISL_622621\|2020 | ···················· | ···················· | ···················· | ···················· | ···················· | ···················· |
| P | 6796 | hCoV-19/Denmark/DCGC-3772/2020\|EPI_ISL_622622\|2020 | ···················· | ···················· | ···················· | ···················· | ···················· | ···················· |
| P | 6797 | hCoV-19/Denmark/DCGC-3777/2020\|EPI_ISL_622623\|2020 | ···················· | ···················· | ···················· | ···················· | ···················· | ···················· |
| P | 6798 | hCoV-19/Denmark/DCGC-3782/2020\|EPI_ISL_622624\|2020 | ···················· | ···················· | ···················· | ···················· | ···················· | ···················· |
| P | 6799 | hCoV-19/Denmark/DCGC-3735/2020\|EPI_ISL_622625\|2020 | ···················· | ···················· | ···················· | ···················· | ···················· | ···················· |
| P | 6800 | hCoV-19/Denmark/DCGC-3740/2020\|EPI_ISL_622626\|2020 | ···················· | ···················· | ···················· | ···················· | ···················· | ···················· |
| P | 6801 | hCoV-19/Denmark/DCGC-3753/2020\|EPI_ISL_622627\|2020 | ···················· | ···················· | ···················· | ···················· | ···················· | ···················· |
| P | 6802 | hCoV-19/Denmark/DCGC-3761/2020\|EPI_ISL_622628\|2020 | ···················· | ···················· | ···················· | ···················· | ···················· | ···················· |
| P | 6803 | hCoV-19/Denmark/DCGC-3768/2020\|EPI_ISL_622629\|2020 | ···················· | ···················· | ···················· | ···················· | ···················· | ···················· |
| P | 6804 | hCoV-19/Denmark/DCGC-3773/2020\|EPI_ISL_622630\|2020 | ···················· | ···················· | ···················· | ···················· | ···················· | ···················· |
| P | 6805 | hCoV-19/Denmark/DCGC-3778/2020\|EPI_ISL_622631\|2020 | ···················· | ···················· | ···················· | ···················· | ···················· | ···················· |
| P | 6806 | hCoV-19/Denmark/DCGC-3736/2020\|EPI_ISL_622633\|2020 | ···················· | ···················· | ···················· | ···················· | ···················· | ···················· |
| P | 6807 | hCoV-19/Denmark/DCGC-3741/2020\|EPI_ISL_622634\|2020 | ···················· | ···················· | ···················· | ···················· | ···················· | ···················· |
| P | 6808 | hCoV-19/Denmark/DCGC-3763/2020\|EPI_ISL_622646\|2020 | ···················· | ···················· | ···················· | ···················· | ···················· | ···················· |
| P | 6809 | hCoV-19/Scotland/QEUH-AAE4FF/2020\|EPI_ISL_623944\|2 | ···················· | ···················· | ···················· | ···················· | ···················· | ···················· |
| P | 6810 | hCoV-19/England/ALDP-AA34FA/2020\|EPI_ISL_623945\|20 | ···················· | ···················· | ···················· | ···················· | ···················· | ···················· |
| P | 6811 | hCoV-19/England/ALDP-AA30C9/2020\|EPI_ISL_623946\|20 | ···················· | ···················· | ···················· | ···················· | ···················· | ···················· |
| P | 6812 | hCoV-19/England/ALDP-AA13FF/2020\|EPI_ISL_623947\|20 | ···················· | ···················· | ···················· | ···················· | ···················· | ···················· |
| P | 6813 | hCoV-19/England/ALDP-AA51D3/2020\|EPI_ISL_623948\|20 | ···················· | ···················· | ···················· | ···················· | ···················· | ···················· |
| P | 6814 | hCoV-19/Scotland/QEUH-AAE5B0/2020\|EPI_ISL_623949\|2 | ···················· | ···················· | ···················· | ···················· | ···················· | ···················· |
| P | 6815 | hCoV-19/England/ALDP-AA0DC5/2020\|EPI_ISL_623950\|20 | ···················· | ···················· | ···················· | ···················· | ···················· | ···················· |
| P | 6816 | hCoV-19/England/ALDP-AA5689/2020\|EPI_ISL_623951\|20 | ···················· | ···················· | ···················· | ···················· | ···················· | ···················· |
| P | 6817 | hCoV-19/England/ALDP-AA2AEA/2020\|EPI_ISL_623952\|20 | ···················· | ·A·················· | ···················· | ···················· | ···················· | ··A················· |
| P | 6818 | hCoV-19/England/ALDP-AA56E3/2020\|EPI_ISL_623953\|20 | ···················· | ·A·················· | ···················· | ···················· | ···················· | ··A················· |
| P | 6819 | hCoV-19/Scotland/QEUH-AAD302/2020\|EPI_ISL_623954\|2 | ···················· | ···················· | ···················· | ···················· | ···················· | ···················· |
| P | 6820 | hCoV-19/England/ALDP-AA19DF/2020\|EPI_ISL_623955\|20 | ···················· | ···················· | ···················· | ···················· | ···················· | ···················· |
| P | 6821 | hCoV-19/England/QEUH-AAD38A/2020\|EPI_ISL_623956\|20 | ···················· | ···················· | ···················· | ···················· | ···················· | ···················· |
| P | 6822 | hCoV-19/England/ALDP-AA12C5/2020\|EPI_ISL_623957\|20 | ···················· | ···················· | ···················· | ···················· | ···················· | ···················· |
| P | 6823 | hCoV-19/England/ALDP-AA2E75/2020\|EPI_ISL_623958\|20 | ···················· | ···················· | ···················· | ···················· | ···················· | ···················· |
| P | 6824 | hCoV-19/England/QEUH-AADC25/2020\|EPI_ISL_623959\|20 | ···················· | ···················· | ···················· | ···················· | ···················· | ···················· |
| P | 6825 | hCoV-19/England/ALDP-AA1395/2020\|EPI_ISL_623961\|20 | ···················· | ···················· | ···················· | ···················· | ······T············· | ···················· |
| P | 6826 | hCoV-19/England/ALDP-AA14B0/2020\|EPI_ISL_623962\|20 | ···················· | ···················· | ···················· | ···················· | ···················· | ···················· |
| P | 6827 | hCoV-19/Scotland/QEUH-AAEF2B/2020\|EPI_ISL_623963\|2 | ···················· | ···················· | ···················· | ···················· | ···················· | ···················· |
| P | 6828 | hCoV-19/England/ALDP-AA516A/2020\|EPI_ISL_623964\|20 | ···················· | ···················· | ···················· | ···················· | ···················· | ···················· |
| P | 6829 | hCoV-19/England/ALDP-AA36D6/2020\|EPI_ISL_623965\|20 | ···················· | ···················· | ···················· | ···················· | ···················· | ···················· |
| P | 6830 | hCoV-19/Scotland/QEUH-AADD04/2020\|EPI_ISL_623977\|2 | ···················· | ···················· | ···················· | ···················· | ···················· | ···················· |
| P | 6831 | hCoV-19/England/ALDP-AA3384/2020\|EPI_ISL_623978\|20 | ···················· | ·A·················· | ···················· | ···················· | ···················· | ··A················· |
| P | 6832 | hCoV-19/Denmark/DCGC-8951/2020\|EPI_ISL_626200\|2020 | ···················· | ···················· | ···················· | ···················· | ···················· | ···················· |
| P | 6833 | hCoV-19/Denmark/DCGC-8962/2020\|EPI_ISL_626201\|2020 | ···················· | ···················· | ···················· | ···················· | ···················· | ···················· |
| P | 6834 | hCoV-19/Denmark/DCGC-8967/2020\|EPI_ISL_626202\|2020 | ···················· | ···················· | ···················· | ···················· | ···················· | ···················· |
| P | 6835 | hCoV-19/Denmark/DCGC-8983/2020\|EPI_ISL_626203\|2020 | ···················· | ···················· | ···················· | ···················· | ···················· | ···················· |
| P | 6836 | hCoV-19/Denmark/DCGC-8996/2020\|EPI_ISL_626204\|2020 | ···················· | ·A·················· | ···················· | ···················· | ···················· | ···················· |
| P | 6837 | hCoV-19/Denmark/DCGC-9004/2020\|EPI_ISL_626205\|2020 | ···················· | ···················· | ···················· | ···················· | ···················· | ···················· |
| P | 6838 | hCoV-19/Denmark/DCGC-9012/2020\|EPI_ISL_626206\|2020 | ···················· | ···················· | ···················· | ···················· | ···················· | ···················· |
| P | 6839 | hCoV-19/Denmark/DCGC-8952/2020\|EPI_ISL_626207\|2020 | ···················· | ···················· | ···················· | ···················· | ···················· | ···················· |
| P | 6840 | hCoV-19/Denmark/DCGC-8968/2020\|EPI_ISL_626208\|2020 | ···················· | ···················· | ···················· | ···················· | ···················· | ···················· |
| P | 6841 | hCoV-19/Denmark/DCGC-8976/2020\|EPI_ISL_626209\|2020 | ···················· | ···················· | ···················· | ···················· | ···················· | ···················· |
| P | 6842 | hCoV-19/Germany/NW-HHU-221/2020\|EPI_ISL_626220\|202 | ···················· | ···················· | ···················· | ···················· | ···················· | ···················· |
| P | 6843 | hCoV-19/Germany/NW-HHU-222/2020\|EPI_ISL_626221\|202 | ···················· | ···················· | ···················· | ···················· | ···················· | ···················· |
| P | 6844 | hCoV-19/Germany/NW-HHU-223/2020\|EPI_ISL_626222\|202 | ···················· | ···················· | ···················· | ···················· | ···················· | ···················· |
| P | 6845 | hCoV-19/Germany/NW-HHU-224/2020\|EPI_ISL_626223\|202 | ···················· | ···················· | ···················· | ···················· | ···················· | ···················· |
| P | 6846 | hCoV-19/Germany/NW-HHU-231/2020\|EPI_ISL_626230\|202 | ···················· | ···················· | ···················· | ···················· | ···················· | ···················· |
| P | 6847 | hCoV-19/Belgium/ULG-10623/2020\|EPI_ISL_626231\|2020 | ···················· | ···················· | ···················· | ···················· | ···················· | ···················· |
| P | 6851 | hCoV-19/Belgium/ULG-10620/2020\|EPI_ISL_626235\|2020 | ···················· | ···················· | ···················· | ···················· | ···················· | ···················· |
| P | 6852 | hCoV-19/Belgium/ULG-10622/2020\|EPI_ISL_626236\|2020 | ···················· | ···················· | ···················· | ···················· | ···················· | ···················· |
| P | 6853 | hCoV-19/Belgium/ULG-10600/2020\|EPI_ISL_626237\|2020 | ···················· | ·A·················· | ···················· | ···················· | ···················· | ···················· |
| P | 6854 | hCoV-19/Belgium/ULG-10601/2020\|EPI_ISL_626238\|2020 | ···················· | ·A·················· | ···················· | ···················· | ···················· | ···················· |
| P | 6856 | hCoV-19/Belgium/ULG-10615/2020\|EPI_ISL_626240\|2020 | ···················· | ···················· | ···················· | ···················· | ···················· | ···················· |
| P | 6862 | hCoV-19/Belgium/ULG-10609/2020\|EPI_ISL_626246\|2020 | ···················· | ···················· | ···················· | ···················· | ···················· | ···················· |
| P | 6863 | hCoV-19/Belgium/ULG-10612/2020\|EPI_ISL_626247\|2020 | ···················· | ···················· | ···················· | ···················· | ···················· | ···················· |
| P | 6864 | hCoV-19/Belgium/ULG-10603/2020\|EPI_ISL_626248\|2020 | ···················· | ···················· | ···················· | ···················· | ···················· | ···················· |
| P | 6865 | hCoV-19/Belgium/ULG-10604/2020\|EPI_ISL_626249\|2020 | ···················· | ···················· | ···················· | ···················· | ···················· | ···················· |
| P | 6866 | hCoV-19/Belgium/ULG-10608/2020\|EPI_ISL_626250\|2020 | ···················· | ·A·················· | ···················· | ···················· | ···················· | ···················· |
| P | 6867 | hCoV-19/Belgium/ULG-10606/2020\|EPI_ISL_626251\|2020 | ···················· | ···················· | ···················· | ···················· | ···················· | ···················· |
| P | 6872 | hCoV-19/Belgium/ULG-10507/2020\|EPI_ISL_626256\|2020 | ···················· | ···················· | ···················· | ···················· | ···················· | ···················· |
| P | 6874 | hCoV-19/Belgium/ULG-10523/2020\|EPI_ISL_626258\|2020 | ···················· | ···················· | ···················· | ···················· | ···················· | ···················· |
| P | 6875 | hCoV-19/Belgium/ULG-10508/2020\|EPI_ISL_626259\|2020 | ···················· | ···················· | ···················· | ···················· | ···················· | ···················· |

| | | | | | | | |
|---|---|---|---|---|---|---|---|
| P | 6876 hCoV-19/Belgium/ULG-10516/2020\|EPI_ISL_626260\|2020 | ···················· | ···················· | ····················· | ···················· | ···················· | ···················· |
| P | 6879 hCoV-19/Belgium/ULG-10520/2020\|EPI_ISL_626263\|2020 | ···················· | ···················· | ····················· | ···················· | ···················· | ···················· |
| P | 6880 hCoV-19/Belgium/ULG-10539/2020\|EPI_ISL_626265\|2020 | ···················· | ···················· | ····················· | ···················· | ···················· | ···················· |
| P | 6884 hCoV-19/Belgium/ULG-10583/2020\|EPI_ISL_626269\|2020 | ···················· | ···················· | ····················· | ···················· | ···················· | ···················· |
| P | 6886 hCoV-19/Belgium/ULG-10510/2020\|EPI_ISL_626271\|2020 | ···················· | ···················· | ····················· | ···················· | ···················· | ···················· |
| P | 6887 hCoV-19/Belgium/ULG-10554/2020\|EPI_ISL_626272\|2020 | ···················· | ···················· | ····················· | ···················· | ···················· | ···················· |
| P | 6888 hCoV-19/Belgium/ULG-10556/2020\|EPI_ISL_626273\|2020 | ···················· | ···················· | ····················· | ···················· | ···················· | ···················· |
| P | 6890 hCoV-19/Belgium/ULG-10568/2020\|EPI_ISL_626275\|2020 | ···················· | ···················· | ····················· | ···················· | ···················· | ···················· |
| P | 6891 hCoV-19/Belgium/ULG-10517/2020\|EPI_ISL_626276\|2020 | ···················· | ···················· | ····················· | ···················· | ···················· | ···················· |
| P | 6893 hCoV-19/Belgium/ULG-10549/2020\|EPI_ISL_626278\|2020 | ···················· | ···················· | ····················· | ···················· | ···················· | ···················· |
| P | 6894 hCoV-19/Belgium/ULG-10521/2020\|EPI_ISL_626279\|2020 | ···················· | ···················· | ····················· | ···················· | ···················· | ···················· |
| P | 6896 hCoV-19/Belgium/ULG-10571/2020\|EPI_ISL_626281\|2020 | ···················· | ···················· | ····················· | ···················· | ···················· | ···················· |
| P | 6897 hCoV-19/Belgium/ULG-10514/2020\|EPI_ISL_626282\|2020 | ···················· | ···················· | ····················· | ···················· | ···················· | ···················· |
| P | 6898 hCoV-19/Belgium/ULG-10558/2020\|EPI_ISL_626283\|2020 | ···················· | ···················· | ····················· | ···················· | ···················· | ···················· |
| P | 6899 hCoV-19/Belgium/ULG-10536/2020\|EPI_ISL_626284\|2020 | ···················· | ···················· | ····················· | ···················· | ···················· | ···················· |
| P | 6900 hCoV-19/Belgium/ULG-10511/2020\|EPI_ISL_626285\|2020 | ···················· | ···················· | ····················· | ···················· | ···················· | ···················· |
| P | 6901 hCoV-19/Belgium/ULG-10540/2020\|EPI_ISL_626286\|2020 | ···················· | ···················· | ····················· | ···················· | ···················· | ···················· |
| P | 6902 hCoV-19/Belgium/ULG-10522/2020\|EPI_ISL_626287\|2020 | ···················· | ···················· | ····················· | ···················· | ···················· | ···················· |
| P | 6903 hCoV-19/Belgium/ULG-10528/2020\|EPI_ISL_626288\|2020 | ···················· | ···················· | ····················· | ···················· | ···················· | ···················· |
| P | 6905 hCoV-19/Belgium/ULG-10546/2020\|EPI_ISL_626290\|2020 | ···················· | ···················· | ····················· | ···················· | ···················· | ···················· |
| P | 6906 hCoV-19/Belgium/ULG-10537/2020\|EPI_ISL_626291\|2020 | ···················· | ···················· | ····················· | ···················· | ···················· | ···················· |
| P | 6907 hCoV-19/Belgium/ULG-10519/2020\|EPI_ISL_626293\|2020 | ···················· | ···················· | ····················· | ···················· | ···················· | ···················· |
| P | 6908 hCoV-19/Belgium/ULG-10590/2020\|EPI_ISL_626294\|2020 | ···················· | ···················· | ····················· | ···················· | ···················· | ···················· |
| P | 6909 hCoV-19/Belgium/ULG-10589/2020\|EPI_ISL_626295\|2020 | ···················· | ···················· | ····················· | ···················· | ···················· | ···················· |
| P | 6911 hCoV-19/Belgium/ULG-10506/2020\|EPI_ISL_626297\|2020 | ···················· | ···················· | ····················· | ···················· | ···················· | ···················· |
| P | 6912 hCoV-19/Belgium/ULG-10541/2020\|EPI_ISL_626299\|2020 | ···················· | ···················· | ····················· | ···················· | ···················· | ···················· |
| P | 6913 hCoV-19/Denmark/DCGC-3023/2020\|EPI_ISL_616801\|2020 | ···················· | ···················· | ····················· | ···················· | ···················· | ···················· |
| P | 6914 hCoV-19/Denmark/DCGC-3025/2020\|EPI_ISL_616803\|2020 | ···················· | ···················· | ····················· | ···················· | ···················· | ···················· |
| P | 6915 hCoV-19/Denmark/DCGC-3027/2020\|EPI_ISL_616805\|2020 | ···················· | ···················· | ····················· | ···················· | ···················· | ···················· |
| P | 6916 hCoV-19/Denmark/DCGC-3028/2020\|EPI_ISL_616806\|2020 | ···················· | ···················· | ····················· | ···················· | ···················· | ···················· |
| P | 6917 hCoV-19/Denmark/DCGC-3029/2020\|EPI_ISL_616807\|2020 | ···················· | ···················· | ····················· | ···················· | ···················· | ···················· |
| P | 6918 hCoV-19/Denmark/DCGC-3031/2020\|EPI_ISL_616809\|2020 | ···················· | ···················· | ····················· | ···················· | ···················· | ···················· |
| P | 6919 hCoV-19/Denmark/DCGC-3032/2020\|EPI_ISL_616810\|2020 | ···················· | ···················· | ····················· | ···················· | ···················· | ···················· |
| P | 6920 hCoV-19/Denmark/DCGC-3033/2020\|EPI_ISL_616811\|2020 | ···················· | ···················· | ····················· | ···················· | ···················· | ···················· |
| P | 6921 hCoV-19/Denmark/DCGC-3034/2020\|EPI_ISL_616812\|2020 | ···················· | ···················· | ····················· | ···················· | ···················· | ···················· |
| P | 6922 hCoV-19/Denmark/DCGC-3035/2020\|EPI_ISL_616813\|2020 | ···················· | ···················· | ····················· | ···················· | ···················· | ···················· |
| P | 6923 hCoV-19/Denmark/DCGC-3036/2020\|EPI_ISL_616814\|2020 | ···················· | ···················· | ····················· | ···················· | ···················· | ···················· |
| P | 6924 hCoV-19/Denmark/DCGC-7195/2020\|EPI_ISL_619000\|2020 | ···················· | ···················· | ····················· | ···················· | ···················· | ···················· |
| P | 6925 hCoV-19/Denmark/DCGC-7196/2020\|EPI_ISL_619001\|2020 | ···················· | ···················· | ····················· | ···················· | ···················· | ···················· |
| P | 6926 hCoV-19/Denmark/DCGC-7204/2020\|EPI_ISL_619002\|2020 | ···················· | ···················· | ····················· | ···················· | ···················· | ···················· |
| P | 6927 hCoV-19/Denmark/DCGC-7208/2020\|EPI_ISL_619003\|2020 | ···················· | ···················· | ····················· | ···················· | ···················· | ···················· |
| P | 6928 hCoV-19/Denmark/DCGC-7217/2020\|EPI_ISL_619004\|2020 | ···················· | ···················· | ····················· | ···················· | ···················· | ···················· |
| P | 6929 hCoV-19/Denmark/DCGC-7221/2020\|EPI_ISL_619005\|2020 | ···················· | ···················· | ····················· | ···················· | ···················· | ···················· |
| P | 6930 hCoV-19/Denmark/DCGC-7223/2020\|EPI_ISL_619006\|2020 | ···················· | ···················· | ····················· | ···················· | ···················· | ···················· |
| P | 6931 hCoV-19/Denmark/DCGC-7225/2020\|EPI_ISL_619007\|2020 | ···················· | ···················· | ····················· | ···················· | ···················· | ···················· |
| P | 6932 hCoV-19/Denmark/DCGC-7226/2020\|EPI_ISL_619008\|2020 | ···················· | ···················· | ····················· | ···················· | ···················· | ···················· |
| P | 6933 hCoV-19/Denmark/DCGC-7227/2020\|EPI_ISL_619009\|2020 | ···················· | ···················· | ····················· | ···················· | ···················· | ···················· |
| P | 6934 hCoV-19/Denmark/DCGC-7247/2020\|EPI_ISL_619010\|2020 | ···················· | ···················· | ····················· | ···················· | ···················· | ···················· |
| P | 6935 hCoV-19/Denmark/DCGC-7251/2020\|EPI_ISL_619011\|2020 | ···················· | ···················· | ····················· | ···················· | ···················· | ···················· |
| P | 6936 hCoV-19/Denmark/DCGC-7262/2020\|EPI_ISL_619012\|2020 | ···················· | ···················· | ····················· | ···················· | ···················· | ···················· |
| P | 6937 hCoV-19/Denmark/DCGC-7263/2020\|EPI_ISL_619013\|2020 | ···················· | ···················· | ····················· | ···················· | ···················· | ···················· |
| P | 6938 hCoV-19/Denmark/DCGC-7265/2020\|EPI_ISL_619014\|2020 | ···················· | ···················· | ····················· | ···················· | ···················· | ···················· |
| P | 6939 hCoV-19/Denmark/DCGC-7287/2020\|EPI_ISL_619015\|2020 | ···················· | ···················· | ····················· | ···················· | ···················· | ···················· |
| P | 6940 hCoV-19/Denmark/DCGC-7290/2020\|EPI_ISL_619016\|2020 | ···················· | ···················· | ····················· | ···················· | ···················· | ···················· |
| P | 6941 hCoV-19/Denmark/DCGC-1616/2020\|EPI_ISL_619017\|2020 | ···················· | ···················· | ····················· | ···················· | ···················· | ···················· |
| P | 6942 hCoV-19/Denmark/DCGC-2048/2020\|EPI_ISL_619018\|2020 | ···················· | ···················· | ····················· | ···················· | ···················· | ···················· |
| P | 6943 hCoV-19/Denmark/DCGC-7320/2020\|EPI_ISL_619019\|2020 | ···················· | ···················· | ····················· | ···················· | ···················· | ···················· |
| P | 6944 hCoV-19/Denmark/DCGC-7357/2020\|EPI_ISL_619027\|2020 | ···················· | ···················· | ····················· | ···················· | ···················· | ···················· |
| P | 6945 hCoV-19/Denmark/DCGC-7465/2020\|EPI_ISL_619050\|2020 | ···················· | ···················· | ····················· | ···················· | ···················· | ···················· |
| P | 6946 hCoV-19/Denmark/DCGC-7511/2020\|EPI_ISL_619051\|2020 | ···················· | ···················· | ····················· | ···················· | ···················· | ···················· |
| P | 6947 hCoV-19/Denmark/DCGC-7154/2020\|EPI_ISL_619053\|2020 | ···················· | ·A·················· | ····················· | ···················· | ···················· | ··A················· |
| P | 6948 hCoV-19/Denmark/DCGC-7201/2020\|EPI_ISL_619054\|2020 | ···················· | ·A·················· | ····················· | ···················· | ···················· | ··A················· |
| P | 6949 hCoV-19/Denmark/DCGC-7348/2020\|EPI_ISL_619055\|2020 | ···················· | ·A·················· | ····················· | ···················· | ···················· | ··A················· |
| P | 6950 hCoV-19/Denmark/DCGC-7350/2020\|EPI_ISL_619056\|2020 | ···················· | ·A·················· | ····················· | ···················· | ···················· | ··A················· |
| P | 6951 hCoV-19/Denmark/DCGC-7351/2020\|EPI_ISL_619057\|2020 | ···················· | ·A·················· | ····················· | ···················· | ···················· | ··A················· |
| P | 6952 hCoV-19/Denmark/DCGC-7305/2020\|EPI_ISL_619058\|2020 | ···················· | ···················· | ····················· | ···················· | ···················· | ···················· |
| P | 6953 hCoV-19/Denmark/DCGC-7436/2020\|EPI_ISL_619059\|2020 | ···················· | ···················· | ····················· | ···················· | ······T············· | ···················· |
| P | 6954 hCoV-19/Denmark/DCGC-7087/2020\|EPI_ISL_619083\|2020 | ···················· | ···················· | ····················· | ···················· | ···················· | ···················· |
| P | 6955 hCoV-19/Denmark/DCGC-7089/2020\|EPI_ISL_619084\|2020 | ···················· | ···················· | ····················· | ···················· | ···················· | ···················· |
| P | 6956 hCoV-19/Denmark/DCGC-7120/2020\|EPI_ISL_619085\|2020 | ···················· | ·A·················· | ····················· | ···················· | ···················· | ···················· |
| P | 6957 hCoV-19/Denmark/DCGC-7185/2020\|EPI_ISL_619086\|2020 | ···················· | ···················· | ····················· | ···················· | ···················· | ···················· |
| P | 6958 hCoV-19/Denmark/DCGC-7186/2020\|EPI_ISL_619087\|2020 | ···················· | ···················· | ····················· | ···················· | ···················· | ···················· |
| P | 6959 hCoV-19/Denmark/DCGC-7222/2020\|EPI_ISL_619088\|2020 | ···················· | ···················· | ····················· | ···················· | ···················· | ···················· |
| P | 6960 hCoV-19/Denmark/DCGC-7311/2020\|EPI_ISL_619089\|2020 | ···················· | ···················· | ····················· | ···················· | ···················· | ···················· |
| P | 6961 hCoV-19/Denmark/DCGC-6246/2020\|EPI_ISL_620006\|2020 | ···················· | ···················· | ····················· | ···················· | ···················· | ···················· |
| P | 6962 hCoV-19/Denmark/DCGC-6250/2020\|EPI_ISL_620010\|2020 | ···················· | ···················· | ····················· | ···················· | ···················· | ···················· |
| P | 6963 hCoV-19/Denmark/DCGC-6251/2020\|EPI_ISL_620011\|2020 | ···················· | ···················· | ····················· | ···················· | ···················· | ···················· |
| P | 6964 hCoV-19/Denmark/DCGC-6252/2020\|EPI_ISL_620012\|2020 | ···················· | ···················· | ····················· | ···················· | ···················· | ···················· |
| P | 6965 hCoV-19/Denmark/DCGC-6253/2020\|EPI_ISL_620013\|2020 | ···················· | ···················· | ····················· | ···················· | ···················· | ···················· |
| P | 6966 hCoV-19/Denmark/DCGC-6254/2020\|EPI_ISL_620014\|2020 | ···················· | ···················· | ····················· | ···················· | ···················· | ···················· |

| | | | | | | | | |
|---|---|---|---|---|---|---|---|---|
| P | 6967 | hCoV-19/Denmark/DCGC-6255/2020\|EPI_ISL_620015\|2020 | ···················· | ···················· | ····················· | ···················· | ···················· | ···················· |
| P | 6968 | hCoV-19/Denmark/DCGC-6256/2020\|EPI_ISL_620016\|2020 | ···················· | ···················· | ····················· | ···················· | ···················· | ···················· |
| P | 6969 | hCoV-19/Denmark/DCGC-6279/2020\|EPI_ISL_620039\|2020 | ···················· | ···················· | ····················· | ···················· | ···················· | ···················· |
| P | 6970 | hCoV-19/Denmark/DCGC-6280/2020\|EPI_ISL_620040\|2020 | ···················· | ···················· | ····················· | ···················· | ···················· | ···················· |
| P | 6971 | hCoV-19/Denmark/DCGC-6285/2020\|EPI_ISL_620045\|2020 | ···················· | ···················· | ····················· | ···················· | ···················· | ···················· |
| P | 6972 | hCoV-19/Denmark/DCGC-6286/2020\|EPI_ISL_620046\|2020 | ···················· | ···················· | ····················· | ···················· | ···················· | ···················· |
| P | 6973 | hCoV-19/Denmark/DCGC-6287/2020\|EPI_ISL_620047\|2020 | ···················· | ···················· | ····················· | ···················· | ···················· | ···················· |
| P | 6974 | hCoV-19/Denmark/DCGC-6288/2020\|EPI_ISL_620048\|2020 | ···················· | ···················· | ····················· | ···················· | ···················· | ···················· |
| P | 6975 | hCoV-19/Denmark/DCGC-6289/2020\|EPI_ISL_620049\|2020 | ···················· | ···················· | ····················· | ···················· | ···················· | ···················· |
| P | 6976 | hCoV-19/Denmark/DCGC-6292/2020\|EPI_ISL_620052\|2020 | ···················· | ···················· | ····················· | ···················· | ···················· | ···················· |
| P | 6977 | hCoV-19/Denmark/DCGC-6294/2020\|EPI_ISL_620054\|2020 | ···················· | ···················· | ····················· | ···················· | ···················· | ···················· |
| P | 6978 | hCoV-19/Denmark/DCGC-6295/2020\|EPI_ISL_620055\|2020 | ···················· | ···················· | ····················· | ···················· | ···················· | ···················· |
| P | 6979 | hCoV-19/Denmark/DCGC-6153/2020\|EPI_ISL_620056\|2020 | ···················· | ···················· | ····················· | ···················· | ···················· | ···················· |
| P | 6980 | hCoV-19/Denmark/DCGC-6155/2020\|EPI_ISL_620058\|2020 | ···················· | ···················· | ····················· | ···················· | ···················· | ···················· |
| P | 6981 | hCoV-19/Denmark/DCGC-6156/2020\|EPI_ISL_620059\|2020 | ···················· | ···················· | ····················· | ···················· | ···················· | ···················· |
| P | 6982 | hCoV-19/Denmark/DCGC-6157/2020\|EPI_ISL_620060\|2020 | ···················· | ···················· | ····················· | ···················· | ···················· | ···················· |
| P | 6983 | hCoV-19/Denmark/DCGC-6158/2020\|EPI_ISL_620061\|2020 | ···················· | ···················· | ····················· | ···················· | ···················· | ···················· |
| P | 6984 | hCoV-19/Denmark/DCGC-6159/2020\|EPI_ISL_620062\|2020 | ···················· | ···················· | ····················· | ···················· | ···················· | ···················· |
| P | 6985 | hCoV-19/Denmark/DCGC-6160/2020\|EPI_ISL_620063\|2020 | ···················· | ···················· | ····················· | ···················· | ···················· | ···················· |
| P | 6986 | hCoV-19/Denmark/DCGC-6161/2020\|EPI_ISL_620064\|2020 | ···················· | ···················· | ····················· | ···················· | ···················· | ···················· |
| P | 6987 | hCoV-19/Denmark/DCGC-6162/2020\|EPI_ISL_620065\|2020 | ···················· | ···················· | ····················· | ···················· | ···················· | ···················· |
| P | 6988 | hCoV-19/Denmark/DCGC-6163/2020\|EPI_ISL_620066\|2020 | ···················· | ···················· | ····················· | ···················· | ···················· | ···················· |
| P | 6989 | hCoV-19/Denmark/DCGC-6164/2020\|EPI_ISL_620067\|2020 | ···················· | ···················· | ····················· | ···················· | ···················· | ···················· |
| P | 6990 | hCoV-19/Denmark/DCGC-6165/2020\|EPI_ISL_620068\|2020 | ···················· | ···················· | ····················· | ···················· | ···················· | ···················· |
| P | 6991 | hCoV-19/Denmark/DCGC-6166/2020\|EPI_ISL_620069\|2020 | ···················· | ···················· | ····················· | ···················· | ···················· | ···················· |
| P | 6992 | hCoV-19/Denmark/DCGC-6167/2020\|EPI_ISL_620070\|2020 | ···················· | ···················· | ····················· | ···················· | ···················· | ···················· |
| P | 6993 | hCoV-19/Denmark/DCGC-6168/2020\|EPI_ISL_620071\|2020 | ···················· | ···················· | ····················· | ···················· | ···················· | ···················· |
| P | 6994 | hCoV-19/Denmark/DCGC-6169/2020\|EPI_ISL_620072\|2020 | ···················· | ···················· | ····················· | ···················· | ···················· | ···················· |
| P | 6995 | hCoV-19/Denmark/DCGC-6170/2020\|EPI_ISL_620073\|2020 | ···················· | ···················· | ····················· | ···················· | ···················· | ···················· |
| P | 6996 | hCoV-19/Denmark/DCGC-6171/2020\|EPI_ISL_620074\|2020 | ···················· | ···················· | ····················· | ···················· | ···················· | ···················· |
| P | 6997 | hCoV-19/Denmark/DCGC-6173/2020\|EPI_ISL_620076\|2020 | ···················· | ···················· | ····················· | ···················· | ···················· | ···················· |
| P | 6998 | hCoV-19/Denmark/DCGC-6175/2020\|EPI_ISL_620078\|2020 | ···················· | ···················· | ····················· | ···················· | ···················· | ···················· |
| P | 6999 | hCoV-19/Denmark/DCGC-6176/2020\|EPI_ISL_620079\|2020 | ···················· | ···················· | ····················· | ···················· | ···················· | ···················· |
| P | 7000 | hCoV-19/Denmark/DCGC-4635/2020\|EPI_ISL_621304\|2020 | ···················· | ···················· | ····················· | ···················· | ···················· | ···················· |
| P | 7001 | hCoV-19/Denmark/DCGC-4636/2020\|EPI_ISL_621305\|2020 | ···················· | ···················· | ····················· | ···················· | ···················· | ···················· |
| P | 7002 | hCoV-19/Denmark/DCGC-4637/2020\|EPI_ISL_621306\|2020 | ···················· | ···················· | ····················· | ···················· | ···················· | ···················· |
| P | 7003 | hCoV-19/Denmark/DCGC-4638/2020\|EPI_ISL_621307\|2020 | ···················· | ···················· | ····················· | ···················· | ···················· | ···················· |
| P | 7004 | hCoV-19/Denmark/DCGC-4639/2020\|EPI_ISL_621308\|2020 | ···················· | ···················· | ····················· | ···················· | ···················· | ···················· |
| P | 7005 | hCoV-19/Denmark/DCGC-4640/2020\|EPI_ISL_621309\|2020 | ···················· | ···················· | ····················· | ···················· | ···················· | ···················· |
| P | 7006 | hCoV-19/Denmark/DCGC-4641/2020\|EPI_ISL_621310\|2020 | ···················· | ···················· | ····················· | ···················· | ···················· | ···················· |
| P | 7007 | hCoV-19/Denmark/DCGC-4516/2020\|EPI_ISL_621312\|2020 | ···················· | ···················· | ····················· | ···················· | ···················· | ···················· |
| P | 7008 | hCoV-19/Denmark/DCGC-4517/2020\|EPI_ISL_621313\|2020 | ···················· | ···················· | ····················· | ···················· | ···················· | ···················· |
| P | 7009 | hCoV-19/Denmark/DCGC-4518/2020\|EPI_ISL_621314\|2020 | ···················· | ···················· | ····················· | ···················· | ···················· | ···················· |
| P | 7010 | hCoV-19/Denmark/DCGC-4524/2020\|EPI_ISL_621320\|2020 | ···················· | ···················· | ····················· | ···················· | ···················· | ···················· |
| P | 7011 | hCoV-19/Denmark/DCGC-4525/2020\|EPI_ISL_621321\|2020 | ···················· | ···················· | ····················· | ···················· | ···················· | ···················· |
| P | 7012 | hCoV-19/Denmark/DCGC-4526/2020\|EPI_ISL_621322\|2020 | ···················· | ···················· | ····················· | ···················· | ···················· | ···················· |
| P | 7013 | hCoV-19/Denmark/DCGC-4527/2020\|EPI_ISL_621323\|2020 | ···················· | ···················· | ····················· | ···················· | ···················· | ···················· |
| P | 7014 | hCoV-19/Denmark/DCGC-4528/2020\|EPI_ISL_621324\|2020 | ···················· | ···················· | ····················· | ···················· | ···················· | ···················· |
| P | 7015 | hCoV-19/Denmark/DCGC-4529/2020\|EPI_ISL_621325\|2020 | ···················· | ···················· | ····················· | ···················· | ···················· | ···················· |
| P | 7016 | hCoV-19/Denmark/DCGC-4530/2020\|EPI_ISL_621326\|2020 | ···················· | ···················· | ····················· | ···················· | ···················· | ···················· |
| P | 7017 | hCoV-19/Denmark/DCGC-4531/2020\|EPI_ISL_621327\|2020 | ···················· | ···················· | ····················· | ···················· | ···················· | ···················· |
| P | 7018 | hCoV-19/Denmark/DCGC-4532/2020\|EPI_ISL_621328\|2020 | ···················· | ···················· | ····················· | ···················· | ···················· | ···················· |
| P | 7019 | hCoV-19/Denmark/DCGC-4533/2020\|EPI_ISL_621329\|2020 | ···················· | ···················· | ····················· | ···················· | ···················· | ···················· |
| P | 7020 | hCoV-19/Denmark/DCGC-4541/2020\|EPI_ISL_621337\|2020 | ···················· | ···················· | ····················· | ···················· | ···················· | ···················· |
| P | 7021 | hCoV-19/Denmark/DCGC-1672/2020\|EPI_ISL_621338\|2020 | ···················· | ···················· | ····················· | ···················· | ···················· | ···················· |
| P | 7022 | hCoV-19/Denmark/DCGC-4542/2020\|EPI_ISL_621339\|2020 | ···················· | ···················· | ····················· | ···················· | ···················· | ···················· |
| P | 7023 | hCoV-19/Denmark/DCGC-4543/2020\|EPI_ISL_621340\|2020 | ···················· | ···················· | ····················· | ···················· | ···················· | ···················· |
| P | 7024 | hCoV-19/Denmark/DCGC-4544/2020\|EPI_ISL_621341\|2020 | ···················· | ···················· | ····················· | ···················· | ···················· | ···················· |
| P | 7025 | hCoV-19/Denmark/DCGC-4545/2020\|EPI_ISL_621342\|2020 | ···················· | ···················· | ····················· | ···················· | ···················· | ···················· |
| P | 7026 | hCoV-19/Denmark/DCGC-4546/2020\|EPI_ISL_621343\|2020 | ···················· | ···················· | ····················· | ···················· | ···················· | ···················· |
| P | 7027 | hCoV-19/Denmark/DCGC-4547/2020\|EPI_ISL_621344\|2020 | ···················· | ···················· | ····················· | ···················· | ···················· | ···················· |
| P | 7028 | hCoV-19/Denmark/DCGC-4548/2020\|EPI_ISL_621345\|2020 | ···················· | ···················· | ····················· | ···················· | ···················· | ···················· |
| P | 7029 | hCoV-19/Denmark/DCGC-4549/2020\|EPI_ISL_621346\|2020 | ···················· | ·A·················· | ····················· | ···················· | ···················· | ··A················· |
| P | 7030 | hCoV-19/Denmark/DCGC-4550/2020\|EPI_ISL_621347\|2020 | ···················· | ···················· | ····················· | ···················· | ···················· | ···················· |
| P | 7031 | hCoV-19/Denmark/DCGC-4573/2020\|EPI_ISL_621370\|2020 | ···················· | ···················· | ····················· | ···················· | ···················· | ···················· |
| P | 7032 | hCoV-19/Denmark/DCGC-4763/2020\|EPI_ISL_621373\|2020 | ···················· | ···················· | ····················· | ···················· | ···················· | ···················· |
| P | 7033 | hCoV-19/Denmark/DCGC-4764/2020\|EPI_ISL_621374\|2020 | ···················· | ···················· | ····················· | ···················· | ···················· | ···················· |
| P | 7034 | hCoV-19/Denmark/DCGC-4765/2020\|EPI_ISL_621375\|2020 | ···················· | ···················· | ····················· | ···················· | ···················· | ···················· |
| P | 7035 | hCoV-19/Denmark/DCGC-4766/2020\|EPI_ISL_621376\|2020 | ···················· | ···················· | ····················· | ···················· | ···················· | ···················· |
| P | 7036 | hCoV-19/Denmark/DCGC-4767/2020\|EPI_ISL_621377\|2020 | ···················· | ···················· | ····················· | ···················· | ···················· | ···················· |
| P | 7037 | hCoV-19/Denmark/DCGC-4768/2020\|EPI_ISL_621378\|2020 | ···················· | ···················· | ····················· | ···················· | ···················· | ···················· |
| P | 7038 | hCoV-19/Denmark/DCGC-4769/2020\|EPI_ISL_621379\|2020 | ···················· | ···················· | ····················· | ···················· | ···················· | ···················· |
| P | 7039 | hCoV-19/Denmark/DCGC-4770/2020\|EPI_ISL_621380\|2020 | ···················· | ···················· | ····················· | ···················· | ···················· | ···················· |
| P | 7040 | hCoV-19/Denmark/DCGC-4771/2020\|EPI_ISL_621381\|2020 | ···················· | ···················· | ····················· | ···················· | ···················· | ···················· |
| P | 7041 | hCoV-19/Denmark/DCGC-4773/2020\|EPI_ISL_621383\|2020 | ···················· | ···················· | ····················· | ···················· | ···················· | ···················· |
| P | 7042 | hCoV-19/Denmark/DCGC-4774/2020\|EPI_ISL_621384\|2020 | ···················· | ···················· | ····················· | ···················· | ···················· | ···················· |
| P | 7043 | hCoV-19/Denmark/DCGC-4775/2020\|EPI_ISL_621385\|2020 | ···················· | ···················· | ····················· | ···················· | ···················· | ···················· |
| P | 7044 | hCoV-19/Denmark/DCGC-4776/2020\|EPI_ISL_621386\|2020 | ···················· | ···················· | ····················· | ···················· | ···················· | ···················· |
| P | 7045 | hCoV-19/Denmark/DCGC-4777/2020\|EPI_ISL_621387\|2020 | ···················· | ···················· | ····················· | ···················· | ···················· | ···················· |
| P | 7046 | hCoV-19/Denmark/DCGC-4778/2020\|EPI_ISL_621388\|2020 | ···················· | ···················· | ····················· | ···················· | ···················· | ···················· |

| | | | | | | | | |
|---|---|---|---|---|---|---|---|---|
| P | 7047 | hCoV-19/Denmark/DCGC-4779/2020\|EPI_ISL_621389\|2020 | .................... | .................... | .................... | .................... | .................... | .................... |
| P | 7048 | hCoV-19/Denmark/DCGC-4780/2020\|EPI_ISL_621390\|2020 | .................... | .................... | .................... | .................... | .................... | .................... |
| P | 7049 | hCoV-19/Denmark/DCGC-4781/2020\|EPI_ISL_621391\|2020 | .................... | .................... | .................... | .................... | .................... | .................... |
| P | 7050 | hCoV-19/Denmark/DCGC-4782/2020\|EPI_ISL_621392\|2020 | .................... | .................... | .................... | .................... | .................... | .................... |
| P | 7051 | hCoV-19/Denmark/DCGC-4783/2020\|EPI_ISL_621393\|2020 | .................... | .................... | .................... | .................... | .................... | .................... |
| P | 7052 | hCoV-19/Denmark/DCGC-4784/2020\|EPI_ISL_621394\|2020 | .................... | .................... | .................... | .................... | .................... | .................... |
| P | 7053 | hCoV-19/Denmark/DCGC-4785/2020\|EPI_ISL_621395\|2020 | .................... | .................... | .................... | .................... | .................... | .................... |
| P | 7054 | hCoV-19/Denmark/DCGC-4786/2020\|EPI_ISL_621396\|2020 | .................... | .................... | .................... | .................... | .................... | .................... |
| P | 7055 | hCoV-19/Denmark/DCGC-4787/2020\|EPI_ISL_621397\|2020 | .................... | .................... | .................... | .................... | .................... | .................... |
| P | 7056 | hCoV-19/Denmark/DCGC-4788/2020\|EPI_ISL_621398\|2020 | .................... | .................... | .................... | .................... | .................... | .................... |
| P | 7057 | hCoV-19/Denmark/DCGC-4789/2020\|EPI_ISL_621399\|2020 | .................... | .................... | .................... | .................... | .................... | .................... |
| P | 7058 | hCoV-19/Denmark/DCGC-1879/2020\|EPI_ISL_622600\|2020 | .................... | .................... | .................... | .................... | .................... | .................... |
| P | 7059 | hCoV-19/Denmark/DCGC-1884/2020\|EPI_ISL_622601\|2020 | .................... | .................... | .................... | .................... | .................... | .................... |
| P | 7060 | hCoV-19/Denmark/DCGC-1892/2020\|EPI_ISL_622602\|2020 | .................... | .................... | .................... | .................... | .................... | .................... |
| P | 7061 | hCoV-19/Denmark/DCGC-1940/2020\|EPI_ISL_622603\|2020 | .................... | .................... | .................... | .................... | .................... | .................... |
| P | 7062 | hCoV-19/Denmark/DCGC-1832/2020\|EPI_ISL_622604\|2020 | .................... | .................... | .................... | .................... | .................... | .................... |
| P | 7063 | hCoV-19/Denmark/DCGC-1840/2020\|EPI_ISL_622605\|2020 | .................... | .................... | .................... | .................... | .................... | .................... |
| P | 7064 | hCoV-19/Denmark/DCGC-1847/2020\|EPI_ISL_622606\|2020 | .................... | .................... | .................... | .................... | .................... | .................... |
| P | 7065 | hCoV-19/Denmark/DCGC-1852/2020\|EPI_ISL_622607\|2020 | .................... | .................... | .................... | .................... | .................... | .................... |
| P | 7066 | hCoV-19/Denmark/DCGC-1860/2020\|EPI_ISL_622608\|2020 | .................... | .A.................. | .................... | .................... | .................... | ..A................. |
| P | 7067 | hCoV-19/Denmark/DCGC-1866/2020\|EPI_ISL_622609\|2020 | .................... | .................... | ..N................. | .................... | .................... | .................... |
| P | 7068 | hCoV-19/Denmark/DCGC-1874/2020\|EPI_ISL_622610\|2020 | .................... | .................... | .................... | .................... | .................... | .................... |
| P | 7069 | hCoV-19/Denmark/DCGC-1885/2020\|EPI_ISL_622612\|2020 | .................... | .................... | .................... | .................... | .................... | .................... |
| P | 7070 | hCoV-19/Denmark/DCGC-3747/2020\|EPI_ISL_622635\|2020 | .................... | .................... | .................... | .................... | .................... | .................... |
| P | 7071 | hCoV-19/Denmark/DCGC-3762/2020\|EPI_ISL_622636\|2020 | .................... | .................... | .................... | .................... | .................... | .................... |
| P | 7072 | hCoV-19/Denmark/DCGC-3769/2020\|EPI_ISL_622637\|2020 | .................... | .................... | .................... | .................... | .................... | .................... |
| P | 7073 | hCoV-19/Denmark/DCGC-3751/2020\|EPI_ISL_622638\|2020 | .................... | .................... | .................... | .................... | .................... | .................... |
| P | 7074 | hCoV-19/Denmark/DCGC-3775/2020\|EPI_ISL_622639\|2020 | .................... | .................... | .................... | .................... | .................... | .................... |
| P | 7075 | hCoV-19/Denmark/DCGC-3779/2020\|EPI_ISL_622640\|2020 | .................... | .................... | .................... | .................... | .................... | .................... |
| P | 7076 | hCoV-19/Denmark/DCGC-3737/2020\|EPI_ISL_622641\|2020 | .................... | .................... | .................... | .................... | .................... | .................... |
| P | 7077 | hCoV-19/Denmark/DCGC-3742/2020\|EPI_ISL_622642\|2020 | .......N............ | .................... | ..........T......... | .................... | .................... | .................... |
| P | 7078 | hCoV-19/Denmark/DCGC-3754/2020\|EPI_ISL_622644\|2020 | .................... | .................... | .................... | .................... | .................... | .................... |
| P | 7079 | hCoV-19/Denmark/DCGC-3758/2020\|EPI_ISL_622645\|2020 | .................... | .................... | .................... | .................... | .................... | .................... |
| P | 7080 | hCoV-19/Denmark/DCGC-3780/2020\|EPI_ISL_622647\|2020 | .................... | .................... | .................... | .................... | .................... | .................... |
| P | 7081 | hCoV-19/Denmark/DCGC-3784/2020\|EPI_ISL_622648\|2020 | .................... | .................... | .................... | .................... | .................... | .................... |
| P | 7082 | hCoV-19/Denmark/DCGC-3743/2020\|EPI_ISL_622650\|2020 | .................... | .................... | .................... | .................... | .................... | .................... |
| P | 7083 | hCoV-19/Denmark/DCGC-3755/2020\|EPI_ISL_622652\|2020 | .................... | .................... | .................... | .................... | .................... | .................... |
| P | 7084 | hCoV-19/Denmark/DCGC-3759/2020\|EPI_ISL_622653\|2020 | .................... | .................... | .................... | .................... | .................... | .................... |
| P | 7085 | hCoV-19/Denmark/DCGC-3765/2020\|EPI_ISL_622654\|2020 | .................... | .................... | .................... | .................... | .................... | .................... |
| P | 7086 | hCoV-19/Denmark/DCGC-3770/2020\|EPI_ISL_622655\|2020 | .................... | .................... | .................... | .................... | .................... | .................... |
| P | 7087 | hCoV-19/Denmark/DCGC-3774/2020\|EPI_ISL_622656\|2020 | .................... | .................... | .................... | .................... | .................... | .................... |
| P | 7088 | hCoV-19/Denmark/DCGC-3776/2020\|EPI_ISL_622657\|2020 | .................... | .................... | .................... | .................... | .................... | .................... |
| P | 7089 | hCoV-19/Denmark/DCGC-3816/2020\|EPI_ISL_622668\|2020 | .................... | .................... | .................... | .................... | .................... | .................... |
| P | 7090 | hCoV-19/Denmark/DCGC-3831/2020\|EPI_ISL_622671\|2020 | .................... | .A..A............... | .................... | .................... | .................... | .................... |
| P | 7091 | hCoV-19/Denmark/DCGC-3841/2020\|EPI_ISL_622673\|2020 | .................... | .................... | .................... | .................... | .................... | .................... |
| P | 7092 | hCoV-19/Denmark/DCGC-3795/2020\|EPI_ISL_622674\|2020 | .................... | .................... | .................... | .................... | .................... | .................... |
| P | 7093 | hCoV-19/Denmark/DCGC-3802/2020\|EPI_ISL_622675\|2020 | .................... | .................... | .................... | .................... | .................... | .................... |
| P | 7094 | hCoV-19/Denmark/DCGC-3810/2020\|EPI_ISL_622677\|2020 | .................... | .................... | .................... | .................... | .................... | .................... |
| P | 7095 | hCoV-19/Denmark/DCGC-3817/2020\|EPI_ISL_622678\|2020 | .................... | .................... | .................... | .................... | .................... | .................... |
| P | 7096 | hCoV-19/England/ALDP-AA2EB1/2020\|EPI_ISL_623900\|20 | .................... | .................... | .................... | .................... | .................... | .................... |
| P | 7097 | hCoV-19/England/ALDP-AA2ACC/2020\|EPI_ISL_623902\|20 | .................... | .................... | .................... | .................... | .................... | .................... |
| P | 7098 | hCoV-19/Scotland/QEUH-AADD7D/2020\|EPI_ISL_623903\|2 | .................... | .................... | .................... | .................... | .................... | .................... |
| P | 7099 | hCoV-19/England/ALDP-AA56C5/2020\|EPI_ISL_623905\|20 | .................... | .................... | .................... | .................... | .................... | .................... |
| P | 7100 | hCoV-19/England/ALDP-AA5276/2020\|EPI_ISL_623906\|20 | .................... | .................... | .................... | .................... | .................... | .................... |
| P | 7101 | hCoV-19/Scotland/QEUH-AADD31/2020\|EPI_ISL_623907\|2 | .................... | .................... | .................... | .................... | .................... | .................... |
| P | 7102 | hCoV-19/England/QEUH-AAD37B/2020\|EPI_ISL_623908\|20 | .................... | .................... | .................... | .................... | .................... | .................... |
| P | 7103 | hCoV-19/England/ALDP-AA126B/2020\|EPI_ISL_623909\|20 | .................... | .................... | .................... | .................... | .................... | .................... |
| P | 7104 | hCoV-19/England/QEUH-AAD706/2020\|EPI_ISL_623911\|20 | .................... | .................... | .................... | .................... | .................... | .................... |
| P | 7105 | hCoV-19/England/ALDP-AA3542/2020\|EPI_ISL_623912\|20 | .................... | .................... | .................... | .................... | .................... | .................... |
| P | 7106 | hCoV-19/England/ALDP-AA32E1/2020\|EPI_ISL_623913\|20 | .................... | .................... | .................... | .................... | .................... | .................... |
| P | 7107 | hCoV-19/England/ALDP-AA11F5/2020\|EPI_ISL_623914\|20 | .................... | .................... | .................... | .................... | .................... | .................... |
| P | 7108 | hCoV-19/England/ALDP-AA2D4B/2020\|EPI_ISL_623915\|20 | .................... | .................... | .................... | .................... | .................... | .................... |
| P | 7109 | hCoV-19/Scotland/QEUH-AAE529/2020\|EPI_ISL_623916\|2 | .................... | .................... | .................... | .................... | .................... | .................... |
| P | 7110 | hCoV-19/England/QEUH-AADBFB/2020\|EPI_ISL_623917\|20 | .................... | .................... | .................... | .................... | ......T............. | .................... |
| P | 7111 | hCoV-19/England/ALDP-AA192A/2020\|EPI_ISL_623918\|20 | .................... | .................... | .................... | .................... | .................... | .................... |
| P | 7112 | hCoV-19/England/ALDP-AA56A7/2020\|EPI_ISL_623919\|20 | .................... | .................... | .................... | .................... | .................... | .................... |
| P | 7113 | hCoV-19/England/ALDP-AA0ED1/2020\|EPI_ISL_623920\|20 | .................... | .................... | .................... | .................... | .................... | .................... |
| P | 7114 | hCoV-19/Scotland/QEUH-AAE5A1/2020\|EPI_ISL_623921\|2 | .................... | .................... | .................... | .................... | .................... | .................... |
| P | 7115 | hCoV-19/England/ALDP-AA1210/2020\|EPI_ISL_623922\|20 | .................... | .................... | .................... | .................... | .................... | .................... |
| P | 7116 | hCoV-19/England/QEUH-AAE4A4/2020\|EPI_ISL_623924\|20 | .................... | .................... | .................... | .................... | .................... | .................... |
| P | 7117 | hCoV-19/England/ALDP-AA40AA/2020\|EPI_ISL_623925\|20 | .................... | .................... | .................... | .................... | .................... | .................... |
| P | 7118 | hCoV-19/England/ALDP-AA2AF9/2020\|EPI_ISL_623927\|20 | .................... | .................... | .................... | .................... | .................... | .................... |
| P | 7119 | hCoV-19/England/ALDP-AA13C2/2020\|EPI_ISL_623928\|20 | .................... | .................... | .................... | .................... | .................... | .................... |
| P | 7120 | hCoV-19/England/ALDP-AA2F63/2020\|EPI_ISL_623929\|20 | .................... | .................... | .................... | .................... | .................... | .................... |
| P | 7121 | hCoV-19/Scotland/QEUH-AADC70/2020\|EPI_ISL_623930\|2 | .................... | .................... | .................... | .................... | .................... | .................... |
| P | 7122 | hCoV-19/England/ALDP-AA2F36/2020\|EPI_ISL_623931\|20 | .................... | .................... | .................... | .................... | .................... | .................... |
| P | 7123 | hCoV-19/England/ALDP-AA330C/2020\|EPI_ISL_623932\|20 | .................... | .A.................. | .................... | .................... | .................... | ..A................. |
| P | 7124 | hCoV-19/Scotland/QEUH-AAE556/2020\|EPI_ISL_623933\|2 | .................... | .................... | .................... | .................... | .................... | .................... |
| P | 7125 | hCoV-19/England/QEUH-AAE4D1/2020\|EPI_ISL_623934\|20 | .................... | .................... | .................... | .................... | .................... | .................... |
| P | 7126 | hCoV-19/England/ALDP-AA1984/2020\|EPI_ISL_623935\|20 | .................... | .................... | .................... | .................... | .................... | .................... |

| | | | | | | | | |
|---|---|---|---|---|---|---|---|---|
| P | 7127 | hCoV-19/England/ALDP-AA11D7/2020\|EPI_ISL_623936\|20 | ···················· | ···················· | ····················· | ···················· | ···················· | ···················· |
| P | 7128 | hCoV-19/England/ALDP-AA13E0/2020\|EPI_ISL_623937\|20 | ···················· | ···················· | ····················· | ···················· | ···················· | ···················· |
| P | 7129 | hCoV-19/England/ALDP-AA374C/2020\|EPI_ISL_623938\|20 | ···················· | ···················· | ····················· | ···················· | ···················· | ···················· |
| P | 7130 | hCoV-19/England/ALDP-AA1429/2020\|EPI_ISL_623940\|20 | ···················· | ·A·················· | ····················· | ···················· | ···················· | ···················· |
| P | 7131 | hCoV-19/England/ALDP-AA318A/2020\|EPI_ISL_623941\|20 | ···················· | ···················· | ····················· | ···················· | ···················· | ···················· |
| P | 7132 | hCoV-19/England/QEUH-AAE477/2020\|EPI_ISL_623942\|20 | ···················· | ···················· | ····················· | ···················· | ···················· | ···················· |
| P | 7133 | hCoV-19/England/ALDP-AA3560/2020\|EPI_ISL_623943\|20 | ···················· | ···················· | ····················· | ···················· | ···················· | ···················· |
| P | 7134 | hCoV-19/England/ALDP-AA078B/2020\|EPI_ISL_623967\|20 | ···················· | ···················· | ····················· | ···················· | ···················· | ···················· |
| P | 7135 | hCoV-19/England/ALDP-AA2DF0/2020\|EPI_ISL_623969\|20 | ···················· | ···················· | ····················· | ···················· | ···················· | ···················· |
| P | 7136 | hCoV-19/England/ALDP-AA5188/2020\|EPI_ISL_623970\|20 | ···················· | ···················· | ····················· | ···················· | ···················· | ···················· |
| P | 7137 | hCoV-19/England/ALDP-AA1975/2020\|EPI_ISL_623971\|20 | ···················· | ···················· | ····················· | ···················· | ···················· | ···················· |
| P | 7138 | hCoV-19/England/ALDP-AA413E/2020\|EPI_ISL_623973\|20 | ···················· | ···················· | ····················· | ···················· | ···················· | ···················· |
| P | 7139 | hCoV-19/Scotland/QEUH-AAE8A8/2020\|EPI_ISL_623975\|2 | ···················· | ···················· | ····················· | ···················· | ···················· | ···················· |
| P | 7140 | hCoV-19/England/QEUH-AAEF67/2020\|EPI_ISL_623976\|20 | ···················· | ···················· | ····················· | ···················· | ···················· | ···················· |
| P | 7141 | hCoV-19/England/ALDP-AA19B1/2020\|EPI_ISL_623981\|20 | ···················· | ···················· | ····················· | ···················· | ···················· | ···················· |
| P | 7142 | hCoV-19/England/ALDP-AA0712/2020\|EPI_ISL_623982\|20 | ···················· | ···················· | ····················· | ···················· | ···················· | ···················· |
| P | 7143 | hCoV-19/England/ALDP-AA06E8/2020\|EPI_ISL_623984\|20 | ···················· | ···················· | ····················· | ···················· | ······T············· | ···················· |
| P | 7144 | hCoV-19/England/ALDP-AA0E2C/2020\|EPI_ISL_623985\|20 | ···················· | ···················· | ····················· | ···················· | ···················· | ···················· |
| P | 7145 | hCoV-19/Scotland/QEUH-AAE583/2020\|EPI_ISL_623986\|2 | ···················· | ···················· | ····················· | ···················· | ···················· | ···················· |
| P | 7146 | hCoV-19/England/ALDP-AA2EDF/2020\|EPI_ISL_623987\|20 | ···················· | ···················· | ····················· | ···················· | ···················· | ···················· |
| P | 7147 | hCoV-19/England/ALDP-AA127A/2020\|EPI_ISL_623988\|20 | ···················· | ···················· | ····················· | ···················· | ···················· | ···················· |
| P | 7148 | hCoV-19/England/ALDP-AA13A4/2020\|EPI_ISL_623999\|20 | ···················· | ···················· | ····················· | ···················· | ···················· | ···················· |
| P | 7149 | hCoV-19/Denmark/DCGC-9008/2020\|EPI_ISL_626180\|2020 | ···················· | ···················· | ····················· | ···················· | ···················· | ···················· |
| P | 7150 | hCoV-19/Denmark/DCGC-8959/2020\|EPI_ISL_626181\|2020 | ···················· | ···················· | ····················· | ···················· | ···················· | ···················· |
| P | 7151 | hCoV-19/Denmark/DCGC-8965/2020\|EPI_ISL_626182\|2020 | ···················· | ···················· | ····················· | ···················· | ···················· | ···················· |
| P | 7152 | hCoV-19/Denmark/DCGC-8972/2020\|EPI_ISL_626183\|2020 | ···················· | ···················· | ····················· | ···················· | ···················· | ···················· |
| P | 7153 | hCoV-19/Denmark/DCGC-8987/2020\|EPI_ISL_626184\|2020 | ···················· | ···················· | ····················· | ···················· | ···················· | ···················· |
| P | 7154 | hCoV-19/Denmark/DCGC-8993/2020\|EPI_ISL_626185\|2020 | ···················· | ···················· | ····················· | ···················· | ···················· | ···················· |
| P | 7155 | hCoV-19/Denmark/DCGC-9001/2020\|EPI_ISL_626186\|2020 | ···················· | ···················· | ····················· | ···················· | ···················· | ···················· |
| P | 7156 | hCoV-19/Denmark/DCGC-9014/2020\|EPI_ISL_626187\|2020 | ···················· | ···················· | ····················· | ···················· | ···················· | ···················· |
| P | 7157 | hCoV-19/Denmark/DCGC-8949/2020\|EPI_ISL_626188\|2020 | ···················· | ···················· | ····················· | ···················· | ···················· | ···················· |
| P | 7158 | hCoV-19/Denmark/DCGC-8960/2020\|EPI_ISL_626189\|2020 | ···················· | ···················· | ····················· | ···················· | ···················· | ···················· |
| P | 7159 | hCoV-19/Denmark/DCGC-8973/2020\|EPI_ISL_626190\|2020 | ···················· | ···················· | ····················· | ···················· | ···················· | ···················· |
| P | 7160 | hCoV-19/Denmark/DCGC-8981/2020\|EPI_ISL_626191\|2020 | ···················· | ···················· | ····················· | ···················· | ···················· | ···················· |
| P | 7161 | hCoV-19/Denmark/DCGC-8988/2020\|EPI_ISL_626192\|2020 | ···················· | ···················· | ····················· | ···················· | ···················· | ···················· |
| P | 7162 | hCoV-19/Denmark/DCGC-9002/2020\|EPI_ISL_626193\|2020 | ···················· | ···················· | ····················· | ···················· | ···················· | ···················· |
| P | 7163 | hCoV-19/Denmark/DCGC-9010/2020\|EPI_ISL_626194\|2020 | ···················· | ···················· | ····················· | ···················· | ···················· | ···················· |
| P | 7164 | hCoV-19/Denmark/DCGC-8950/2020\|EPI_ISL_626195\|2020 | ···················· | ·A·················· | ····················· | ···················· | ···················· | ···················· |
| P | 7165 | hCoV-19/Denmark/DCGC-8953/2020\|EPI_ISL_626196\|2020 | ···················· | ···················· | ····················· | ···················· | ···················· | ···················· |
| P | 7166 | hCoV-19/Denmark/DCGC-8961/2020\|EPI_ISL_626197\|2020 | ···················· | ···················· | ····················· | ···················· | ···················· | ···················· |
| P | 7167 | hCoV-19/Denmark/DCGC-8974/2020\|EPI_ISL_626198\|2020 | ···················· | ···················· | ····················· | ···················· | ···················· | ···················· |
| P | 7168 | hCoV-19/Denmark/DCGC-8989/2020\|EPI_ISL_626199\|2020 | ···················· | ···················· | ····················· | ···················· | ···················· | ···················· |
| P | 7169 | hCoV-19/Denmark/DCGC-7636/2020\|EPI_ISL_618520\|2020 | ···················· | ···················· | ····················· | ···················· | ···················· | ···················· |
| P | 7170 | hCoV-19/Denmark/DCGC-7653/2020\|EPI_ISL_618522\|2020 | ···················· | ···················· | ····················· | ···················· | ···················· | ···················· |
| P | 7171 | hCoV-19/Denmark/DCGC-7612/2020\|EPI_ISL_618524\|2020 | ···················· | ·A·················· | ····················· | ···················· | ···················· | ···················· |
| P | 7172 | hCoV-19/Denmark/DCGC-1579/2020\|EPI_ISL_618525\|2020 | ···················· | ···················· | ····················· | ···················· | ···················· | ···················· |
| P | 7173 | hCoV-19/Denmark/DCGC-7620/2020\|EPI_ISL_618526\|2020 | ···················· | ···················· | ····················· | ···················· | ···················· | ···················· |
| P | 7174 | hCoV-19/Denmark/DCGC-7626/2020\|EPI_ISL_618527\|2020 | ···················· | ···················· | ····················· | ···················· | ···················· | ···················· |
| P | 7175 | hCoV-19/Denmark/DCGC-7637/2020\|EPI_ISL_618528\|2020 | ···················· | ···················· | ····················· | ···················· | ···················· | ···················· |
| P | 7176 | hCoV-19/Denmark/DCGC-7326/2020\|EPI_ISL_619020\|2020 | ···················· | ···················· | ····················· | ···················· | ···················· | ···················· |
| P | 7177 | hCoV-19/Denmark/DCGC-7321/2020\|EPI_ISL_619021\|2020 | ···················· | ···················· | ····················· | ···················· | ···················· | ···················· |
| P | 7178 | hCoV-19/Denmark/DCGC-7355/2020\|EPI_ISL_619022\|2020 | ···················· | ···················· | ····················· | ···················· | ···················· | ···················· |
| P | 7179 | hCoV-19/Denmark/DCGC-7377/2020\|EPI_ISL_619023\|2020 | ···················· | ···················· | ····················· | ···················· | ···················· | ···················· |
| P | 7180 | hCoV-19/Denmark/DCGC-7356/2020\|EPI_ISL_619024\|2020 | ···················· | ···················· | ····················· | ···················· | ···················· | ···················· |
| P | 7181 | hCoV-19/Denmark/DCGC-7363/2020\|EPI_ISL_619025\|2020 | ···················· | ···················· | ····················· | ···················· | ···················· | ···················· |
| P | 7182 | hCoV-19/Denmark/DCGC-7371/2020\|EPI_ISL_619026\|2020 | ···················· | ···················· | ····················· | ···················· | ···················· | ···················· |
| P | 7183 | hCoV-19/Denmark/DCGC-7372/2020\|EPI_ISL_619028\|2020 | ···················· | ···················· | ····················· | ···················· | ···················· | ···················· |
| P | 7184 | hCoV-19/Denmark/DCGC-7338/2020\|EPI_ISL_619029\|2020 | ···················· | ···················· | ····················· | ···················· | ···················· | ···················· |
| P | 7185 | hCoV-19/Denmark/DCGC-7367/2020\|EPI_ISL_619030\|2020 | ···················· | ···················· | ····················· | ···················· | ···················· | ···················· |
| P | 7186 | hCoV-19/Denmark/DCGC-7374/2020\|EPI_ISL_619031\|2020 | ···················· | ···················· | ····················· | ···················· | ···················· | ···················· |
| P | 7187 | hCoV-19/Denmark/DCGC-7380/2020\|EPI_ISL_619032\|2020 | ···················· | ···················· | ····················· | ···················· | ···················· | ···················· |
| P | 7188 | hCoV-19/Denmark/DCGC-7398/2020\|EPI_ISL_619033\|2020 | ···················· | ···················· | ····················· | ···················· | ···················· | ···················· |
| P | 7189 | hCoV-19/Denmark/DCGC-7381/2020\|EPI_ISL_619034\|2020 | ···················· | ···················· | ····················· | ···················· | ···················· | ···················· |
| P | 7190 | hCoV-19/Denmark/DCGC-7387/2020\|EPI_ISL_619035\|2020 | ···················· | ···················· | ····················· | ···················· | ···················· | ···················· |
| P | 7191 | hCoV-19/Denmark/DCGC-7404/2020\|EPI_ISL_619036\|2020 | ···················· | ···················· | ····················· | ···················· | ···················· | ···················· |
| P | 7192 | hCoV-19/Denmark/DCGC-7435/2020\|EPI_ISL_619037\|2020 | ···················· | ···················· | ····················· | ···················· | ···················· | ···················· |
| P | 7193 | hCoV-19/Denmark/DCGC-7451/2020\|EPI_ISL_619038\|2020 | ···················· | ···················· | ····················· | ···················· | ···················· | ···················· |
| P | 7194 | hCoV-19/Denmark/DCGC-7382/2020\|EPI_ISL_619039\|2020 | ···················· | ···················· | ····················· | ···················· | ···················· | ···················· |
| P | 7195 | hCoV-19/Denmark/DCGC-7405/2020\|EPI_ISL_619040\|2020 | ···················· | ···················· | ····················· | ···················· | ···················· | ···················· |
| P | 7196 | hCoV-19/Denmark/DCGC-7453/2020\|EPI_ISL_619041\|2020 | ···················· | ···················· | ····················· | ···················· | ···················· | ···················· |
| P | 7197 | hCoV-19/Denmark/DCGC-7407/2020\|EPI_ISL_619042\|2020 | ···················· | ···················· | ····················· | ···················· | ···················· | ···················· |
| P | 7198 | hCoV-19/Denmark/DCGC-7454/2020\|EPI_ISL_619043\|2020 | ···················· | ···················· | ····················· | ···················· | ···················· | ···················· |
| P | 7199 | hCoV-19/Denmark/DCGC-7450/2020\|EPI_ISL_619044\|2020 | ···················· | ···················· | ····················· | ···················· | ···················· | ···················· |
| P | 7200 | hCoV-19/Denmark/DCGC-7462/2020\|EPI_ISL_619045\|2020 | ···················· | ···················· | ····················· | ···················· | ···················· | ···················· |
| P | 7201 | hCoV-19/Denmark/DCGC-7517/2020\|EPI_ISL_619046\|2020 | ···················· | ···················· | ····················· | ···················· | ···················· | ···················· |
| P | 7202 | hCoV-19/Denmark/DCGC-7464/2020\|EPI_ISL_619047\|2020 | ···················· | ···················· | ····················· | ···················· | ···················· | ···················· |
| P | 7203 | hCoV-19/Denmark/DCGC-7518/2020\|EPI_ISL_619048\|2020 | ···················· | ···················· | ····················· | ···················· | ···················· | ···················· |
| P | 7204 | hCoV-19/Denmark/DCGC-7524/2020\|EPI_ISL_619049\|2020 | ···················· | ···················· | ····················· | ···················· | ···················· | ···················· |
| P | 7205 | hCoV-19/Denmark/DCGC-6954/2020\|EPI_ISL_619800\|2020 | ···················· | ···················· | ····················· | ···················· | ···················· | ···················· |
| P | 7206 | hCoV-19/Denmark/DCGC-6955/2020\|EPI_ISL_619801\|2020 | ···················· | ···················· | ····················· | ···················· | ···················· | ···················· |

| | | | | | | | |
|---|---|---|---|---|---|---|---|
| P | 7207 hCoV-19/Denmark/DCGC-6956/2020\|EPI_ISL_619802\|2020 | ···················· | ·A·················· | ···················· | ···················· | ···················· | ···················· |
| P | 7208 hCoV-19/Denmark/DCGC-6957/2020\|EPI_ISL_619803\|2020 | ···················· | ···················· | ···················· | ···················· | ···················· | ···················· |
| P | 7209 hCoV-19/Denmark/DCGC-6958/2020\|EPI_ISL_619804\|2020 | ···················· | ·A·················· | ···················· | ···················· | ···················· | ···················· |
| P | 7210 hCoV-19/Denmark/DCGC-6959/2020\|EPI_ISL_619805\|2020 | ···················· | ···················· | ···················· | ···················· | ···················· | ···················· |
| P | 7211 hCoV-19/Denmark/DCGC-6960/2020\|EPI_ISL_619806\|2020 | ···················· | ···················· | ···················· | ···················· | ···················· | ···················· |
| P | 7212 hCoV-19/Denmark/DCGC-7004/2020\|EPI_ISL_619851\|2020 | ···················· | ···················· | ···················· | ···················· | ···················· | ···················· |
| P | 7213 hCoV-19/Denmark/DCGC-7006/2020\|EPI_ISL_619853\|2020 | ···················· | ···················· | ···················· | ···················· | ···················· | ···················· |
| P | 7214 hCoV-19/Denmark/DCGC-7007/2020\|EPI_ISL_619854\|2020 | ···················· | ···················· | ···················· | ···················· | ···················· | ···················· |
| P | 7215 hCoV-19/Denmark/DCGC-7008/2020\|EPI_ISL_619855\|2020 | ···················· | ···················· | ···················· | ···················· | ···················· | ···················· |
| P | 7216 hCoV-19/Denmark/DCGC-7009/2020\|EPI_ISL_619856\|2020 | ···················· | ···················· | ···················· | ···················· | ···················· | ···················· |
| P | 7217 hCoV-19/Denmark/DCGC-7010/2020\|EPI_ISL_619857\|2020 | ···················· | ···················· | ···················· | ···················· | ···················· | ···················· |
| P | 7218 hCoV-19/Denmark/DCGC-7011/2020\|EPI_ISL_619858\|2020 | ···················· | ···················· | ···················· | ···················· | ···················· | ···················· |
| P | 7219 hCoV-19/Denmark/DCGC-7012/2020\|EPI_ISL_619859\|2020 | ···················· | ···················· | ···················· | ···················· | ···················· | ···················· |
| P | 7220 hCoV-19/Denmark/DCGC-6240/2020\|EPI_ISL_620000\|2020 | ···················· | ···················· | ···················· | ···················· | ···················· | ···················· |
| P | 7221 hCoV-19/Denmark/DCGC-6241/2020\|EPI_ISL_620001\|2020 | ···················· | ···················· | ···················· | ···················· | ···················· | ···················· |
| P | 7222 hCoV-19/Denmark/DCGC-6243/2020\|EPI_ISL_620003\|2020 | ···················· | ···················· | ···················· | ···················· | ···················· | ···················· |
| P | 7223 hCoV-19/Denmark/DCGC-6244/2020\|EPI_ISL_620004\|2020 | ···················· | ···················· | ···················· | ···················· | ···················· | ···················· |
| P | 7224 hCoV-19/Denmark/DCGC-6257/2020\|EPI_ISL_620017\|2020 | ···················· | ···················· | ···················· | ···················· | ···················· | ···················· |
| P | 7225 hCoV-19/Denmark/DCGC-6258/2020\|EPI_ISL_620018\|2020 | ···················· | ···················· | ···················· | ···················· | ···················· | ···················· |
| P | 7226 hCoV-19/Denmark/DCGC-6259/2020\|EPI_ISL_620019\|2020 | ···················· | ···················· | ···················· | ···················· | ···················· | ···················· |
| P | 7227 hCoV-19/Denmark/DCGC-6260/2020\|EPI_ISL_620020\|2020 | ···················· | ···················· | ···················· | ···················· | ···················· | ···················· |
| P | 7228 hCoV-19/Denmark/DCGC-6261/2020\|EPI_ISL_620021\|2020 | ···················· | ···················· | ···················· | ···················· | ···················· | ···················· |
| P | 7229 hCoV-19/Denmark/DCGC-6262/2020\|EPI_ISL_620022\|2020 | ···················· | ···················· | ···················· | ···················· | ···················· | ···················· |
| P | 7230 hCoV-19/Denmark/DCGC-6263/2020\|EPI_ISL_620023\|2020 | ···················· | ···················· | ···················· | ···················· | ···················· | ···················· |
| P | 7231 hCoV-19/Denmark/DCGC-6264/2020\|EPI_ISL_620024\|2020 | ···················· | ···················· | ···················· | ···················· | ···················· | ···················· |
| P | 7232 hCoV-19/Denmark/DCGC-6265/2020\|EPI_ISL_620025\|2020 | ···················· | ···················· | ···················· | ···················· | ···················· | ···················· |
| P | 7233 hCoV-19/Denmark/DCGC-6266/2020\|EPI_ISL_620026\|2020 | ···················· | ···················· | ···················· | ···················· | ···················· | ···················· |
| P | 7234 hCoV-19/Denmark/DCGC-6267/2020\|EPI_ISL_620027\|2020 | ···················· | ·A·················· | ···················· | ···················· | ···················· | ··A················· |
| P | 7235 hCoV-19/Denmark/DCGC-6268/2020\|EPI_ISL_620028\|2020 | ···················· | ···················· | ···················· | ···················· | ···················· | ···················· |
| P | 7236 hCoV-19/Denmark/DCGC-6269/2020\|EPI_ISL_620029\|2020 | ···················· | ···················· | ···················· | ···················· | ···················· | ···················· |
| P | 7237 hCoV-19/Denmark/DCGC-6270/2020\|EPI_ISL_620030\|2020 | ···················· | ···················· | ···················· | ···················· | ···················· | ···················· |
| P | 7238 hCoV-19/Denmark/DCGC-6272/2020\|EPI_ISL_620032\|2020 | ···················· | ···················· | ···················· | ···················· | ···················· | ···················· |
| P | 7239 hCoV-19/Denmark/DCGC-6273/2020\|EPI_ISL_620033\|2020 | ···················· | ···················· | ···················· | ···················· | ···················· | ···················· |
| P | 7240 hCoV-19/Denmark/DCGC-6274/2020\|EPI_ISL_620034\|2020 | ···················· | ···················· | ···················· | ···················· | ···················· | ···················· |
| P | 7241 hCoV-19/Denmark/DCGC-6275/2020\|EPI_ISL_620035\|2020 | ···················· | ···················· | ···················· | ···················· | ···················· | ···················· |
| P | 7242 hCoV-19/Denmark/DCGC-6276/2020\|EPI_ISL_620036\|2020 | ···················· | ···················· | ···················· | ···················· | ···················· | ···················· |
| P | 7243 hCoV-19/Denmark/DCGC-6277/2020\|EPI_ISL_620037\|2020 | ···················· | ···················· | ···················· | ···················· | ···················· | ···················· |
| P | 7244 hCoV-19/Denmark/DCGC-6278/2020\|EPI_ISL_620038\|2020 | ···················· | ···················· | ···················· | ···················· | ···················· | ···················· |
| P | 7245 hCoV-19/Denmark/DCGC-5517/2020\|EPI_ISL_620842\|2020 | ···················· | ···················· | ···················· | ···················· | ···················· | ···················· |
| P | 7246 hCoV-19/Denmark/DCGC-5518/2020\|EPI_ISL_620843\|2020 | ···················· | ···················· | ···················· | ···················· | ···················· | ···················· |
| P | 7247 hCoV-19/Denmark/DCGC-5520/2020\|EPI_ISL_620845\|2020 | ···················· | ···················· | ···················· | ···················· | ···················· | ···················· |
| P | 7248 hCoV-19/Denmark/DCGC-5521/2020\|EPI_ISL_620846\|2020 | ···················· | ···················· | ···················· | ···················· | ···················· | ···················· |
| P | 7249 hCoV-19/Denmark/DCGC-5522/2020\|EPI_ISL_620847\|2020 | ···················· | ···················· | ···················· | ···················· | ···················· | ···················· |
| P | 7250 hCoV-19/Denmark/DCGC-5523/2020\|EPI_ISL_620848\|2020 | ···················· | ···················· | ···················· | ···················· | ···················· | ···················· |
| P | 7251 hCoV-19/Denmark/DCGC-5524/2020\|EPI_ISL_620849\|2020 | ···················· | ···················· | ···················· | ···················· | ···················· | ···················· |
| P | 7252 hCoV-19/Denmark/DCGC-4534/2020\|EPI_ISL_621330\|2020 | ···················· | ···················· | ···················· | ···················· | ···················· | ···················· |
| P | 7253 hCoV-19/Denmark/DCGC-4535/2020\|EPI_ISL_621331\|2020 | ···················· | ···················· | ···················· | ···················· | ···················· | ···················· |
| P | 7254 hCoV-19/Denmark/DCGC-4536/2020\|EPI_ISL_621332\|2020 | ···················· | ···················· | ···················· | ···················· | ···················· | ···················· |
| P | 7255 hCoV-19/Denmark/DCGC-4537/2020\|EPI_ISL_621333\|2020 | ···················· | ···················· | ···················· | ···················· | ···················· | ···················· |
| P | 7256 hCoV-19/Denmark/DCGC-4538/2020\|EPI_ISL_621334\|2020 | ···················· | ···················· | ···················· | ···················· | ···················· | ···················· |
| P | 7257 hCoV-19/Denmark/DCGC-4539/2020\|EPI_ISL_621335\|2020 | ···················· | ···················· | ···················· | ···················· | ···················· | ···················· |
| P | 7258 hCoV-19/Denmark/DCGC-4540/2020\|EPI_ISL_621336\|2020 | ···················· | ···················· | ···················· | ···················· | ···················· | ···················· |
| P | 7259 hCoV-19/Denmark/DCGC-4551/2020\|EPI_ISL_621348\|2020 | ···················· | ···················· | ···················· | ···················· | ···················· | ···················· |
| P | 7260 hCoV-19/Denmark/DCGC-4553/2020\|EPI_ISL_621350\|2020 | ···················· | ···················· | ···················· | ···················· | ···················· | ···················· |
| P | 7261 hCoV-19/Denmark/DCGC-4554/2020\|EPI_ISL_621351\|2020 | ···················· | ···················· | ···················· | ···················· | ···················· | ···················· |
| P | 7262 hCoV-19/Denmark/DCGC-4555/2020\|EPI_ISL_621352\|2020 | ···················· | ···················· | ···················· | ···················· | ···················· | ···················· |
| P | 7263 hCoV-19/Denmark/DCGC-4556/2020\|EPI_ISL_621353\|2020 | ···················· | ···················· | ···················· | ···················· | ···················· | ···················· |
| P | 7264 hCoV-19/Denmark/DCGC-4557/2020\|EPI_ISL_621354\|2020 | ···················· | ···················· | ···················· | ···················· | ···················· | ···················· |
| P | 7265 hCoV-19/Denmark/DCGC-4558/2020\|EPI_ISL_621355\|2020 | ···················· | ···················· | ···················· | ···················· | ···················· | ···················· |
| P | 7266 hCoV-19/Denmark/DCGC-4559/2020\|EPI_ISL_621356\|2020 | ···················· | ···················· | ···················· | ···················· | ···················· | ···················· |
| P | 7267 hCoV-19/Denmark/DCGC-4560/2020\|EPI_ISL_621357\|2020 | ···················· | ···················· | ···················· | ···················· | ···················· | ···················· |
| P | 7268 hCoV-19/Denmark/DCGC-4561/2020\|EPI_ISL_621358\|2020 | ···················· | ···················· | ···················· | ···················· | ···················· | ···················· |
| P | 7269 hCoV-19/Denmark/DCGC-4562/2020\|EPI_ISL_621359\|2020 | ···················· | ···················· | ···················· | ···················· | ···················· | ···················· |
| P | 7270 hCoV-19/Denmark/DCGC-4563/2020\|EPI_ISL_621360\|2020 | ···················· | ···················· | ···················· | ···················· | ···················· | ···················· |
| P | 7271 hCoV-19/Denmark/DCGC-4564/2020\|EPI_ISL_621361\|2020 | ···················· | ···················· | ···················· | ···················· | ···················· | ···················· |
| P | 7272 hCoV-19/Denmark/DCGC-4565/2020\|EPI_ISL_621362\|2020 | ···················· | ···················· | ···················· | ···················· | ···················· | ···················· |
| P | 7273 hCoV-19/Denmark/DCGC-4566/2020\|EPI_ISL_621363\|2020 | ···················· | ···················· | ···················· | ···················· | ···················· | ···················· |
| P | 7274 hCoV-19/Denmark/DCGC-4567/2020\|EPI_ISL_621364\|2020 | ···················· | ···················· | ···················· | ···················· | ···················· | ···················· |
| P | 7275 hCoV-19/Denmark/DCGC-4568/2020\|EPI_ISL_621365\|2020 | ···················· | ···················· | ···················· | ···················· | ···················· | ···················· |
| P | 7276 hCoV-19/Denmark/DCGC-4569/2020\|EPI_ISL_621366\|2020 | ···················· | ···················· | ···················· | ···················· | ···················· | ···················· |
| P | 7277 hCoV-19/Denmark/DCGC-4570/2020\|EPI_ISL_621367\|2020 | ···················· | ···················· | ···················· | ···················· | ···················· | ···················· |
| P | 7278 hCoV-19/Denmark/DCGC-4572/2020\|EPI_ISL_621369\|2020 | ···················· | ···················· | ···················· | ···················· | ···················· | ···················· |
| P | 7279 hCoV-19/Denmark/DCGC-3744/2020\|EPI_ISL_622658\|2020 | ···················· | ···················· | ···················· | ···················· | ···················· | ···················· |
| P | 7280 hCoV-19/Denmark/DCGC-3750/2020\|EPI_ISL_622659\|2020 | ···················· | ···················· | ···················· | ···················· | ···················· | ···················· |
| P | 7281 hCoV-19/Denmark/DCGC-3745/2020\|EPI_ISL_622660\|2020 | ···················· | ···················· | ···················· | ···················· | ···················· | ···················· |
| P | 7282 hCoV-19/Denmark/DCGC-3764/2020\|EPI_ISL_622661\|2020 | ···················· | ···················· | ···················· | ···················· | ···················· | ···················· |
| P | 7283 hCoV-19/Denmark/DCGC-3766/2020\|EPI_ISL_622662\|2020 | ···················· | ···················· | ···················· | ···················· | ···················· | ···················· |
| P | 7284 hCoV-19/Denmark/DCGC-3771/2020\|EPI_ISL_622663\|2020 | ···················· | ···················· | ···················· | ···················· | ···················· | ···················· |
| P | 7285 hCoV-19/Denmark/DCGC-3756/2020\|EPI_ISL_622664\|2020 | ···················· | ···················· | ···················· | ···················· | ···················· | ···················· |
| P | 7286 hCoV-19/Denmark/DCGC-3781/2020\|EPI_ISL_622665\|2020 | ···················· | ···················· | ···················· | ···················· | ···················· | ···················· |

| | | | | | | | |
|---|---|---|---|---|---|---|---|
| P | 7287 hCoV-19/Denmark/DCGC-3794/2020\|EPI_ISL_622666\|2020 | ···················· | ·A·················· | ···················· | ···················· | ···················· | ··A················· |
| P | 7288 hCoV-19/Denmark/DCGC-3801/2020\|EPI_ISL_622667\|2020 | ···················· | ···················· | ···················· | ···················· | ···················· | ···················· |
| P | 7289 hCoV-19/Denmark/DCGC-1651/2020\|EPI_ISL_622679\|2020 | ···················· | ···················· | ···················· | ···················· | ···················· | ···················· |
| P | 7290 hCoV-19/Denmark/DCGC-3822/2020\|EPI_ISL_622680\|2020 | ···················· | ·A·················· | ···················· | ···················· | ···················· | ··A················· |
| P | 7291 hCoV-19/Denmark/DCGC-3827/2020\|EPI_ISL_622681\|2020 | ···················· | ·A·················· | ···················· | ···················· | ···················· | ··A················· |
| P | 7292 hCoV-19/Denmark/DCGC-3785/2020\|EPI_ISL_622682\|2020 | ···················· | ···················· | ···················· | ···················· | ···················· | ···················· |
| P | 7293 hCoV-19/Denmark/DCGC-3791/2020\|EPI_ISL_622683\|2020 | ···················· | ···················· | ···················· | ···················· | ···················· | ···················· |
| P | 7294 hCoV-19/Denmark/DCGC-3796/2020\|EPI_ISL_622684\|2020 | ···················· | ···················· | ···················· | ···················· | ···················· | ···················· |
| P | 7295 hCoV-19/Denmark/DCGC-3790/2020\|EPI_ISL_622685\|2020 | ···················· | ···················· | ···················· | ···················· | ······T············· | ···················· |
| P | 7296 hCoV-19/Denmark/DCGC-3811/2020\|EPI_ISL_622686\|2020 | ···················· | ···················· | ···················· | ···················· | ······T············· | ···················· |
| P | 7297 hCoV-19/Denmark/DCGC-3832/2020\|EPI_ISL_622687\|2020 | ···················· | ···················· | ···················· | ···················· | ···················· | ···················· |
| P | 7298 hCoV-19/Denmark/DCGC-3837/2020\|EPI_ISL_622688\|2020 | ···················· | ···················· | ···················· | ···················· | ···················· | ···················· |
| P | 7299 hCoV-19/Denmark/DCGC-3786/2020\|EPI_ISL_622689\|2020 | ···················· | ···················· | ···················· | ···················· | ···················· | ···················· |
| P | 7300 hCoV-19/Denmark/DCGC-3797/2020\|EPI_ISL_622690\|2020 | ···················· | ···················· | ···················· | ···················· | ···················· | ···················· |
| P | 7301 hCoV-19/Denmark/DCGC-3803/2020\|EPI_ISL_622691\|2020 | ···················· | ···················· | ···················· | ···················· | ···················· | ···················· |
| P | 7302 hCoV-19/Denmark/DCGC-3807/2020\|EPI_ISL_622692\|2020 | ···················· | ···················· | ···················· | ···················· | ···················· | ···················· |
| P | 7303 hCoV-19/Denmark/DCGC-3812/2020\|EPI_ISL_622693\|2020 | ···················· | ···················· | ···················· | ···················· | ···················· | ···················· |
| P | 7304 hCoV-19/Denmark/DCGC-3823/2020\|EPI_ISL_622695\|2020 | ···················· | ···················· | ···················· | ···················· | ···················· | ···················· |
| P | 7305 hCoV-19/Denmark/DCGC-3828/2020\|EPI_ISL_622696\|2020 | ···················· | ···················· | ···················· | ···················· | ···················· | ···················· |
| P | 7306 hCoV-19/Denmark/DCGC-3833/2020\|EPI_ISL_622697\|2020 | ···················· | ···················· | ···················· | ···················· | ···················· | ···················· |
| P | 7307 hCoV-19/Denmark/DCGC-3842/2020\|EPI_ISL_622698\|2020 | ···················· | ···················· | ···················· | ···················· | ···················· | ···················· |
| P | 7308 hCoV-19/Denmark/DCGC-3787/2020\|EPI_ISL_622699\|2020 | ···················· | ···················· | ···················· | ···················· | ···················· | ···················· |
| P | 7309 hCoV-19/England/ALDP-AA40E6/2020\|EPI_ISL_623989\|20 | ···················· | ···················· | ···················· | ···················· | ···················· | ···················· |
| P | 7310 hCoV-19/Scotland/QEUH-AAE547/2020\|EPI_ISL_623990\|2 | ···················· | ···················· | ···················· | ···················· | ···················· | ···················· |
| P | 7311 hCoV-19/England/QEUH-AAD399/2020\|EPI_ISL_623991\|20 | ···················· | ···················· | ···················· | ···················· | ···················· | ···················· |
| P | 7312 hCoV-19/England/ALDP-AA16AB/2020\|EPI_ISL_623992\|20 | ···················· | ···················· | ···················· | ···················· | ···················· | ···················· |
| P | 7313 hCoV-19/England/ALDP-AA11E6/2020\|EPI_ISL_623993\|20 | ···················· | ···················· | ···················· | ···················· | ···················· | ···················· |
| P | 7314 hCoV-19/Scotland/QEUH-AADC52/2020\|EPI_ISL_623994\|2 | ···················· | ···················· | ···················· | ···················· | ···················· | ···················· |
| P | 7315 hCoV-19/Scotland/QEUH-AAEEC4/2020\|EPI_ISL_623995\|2 | ···················· | ···················· | ···················· | ···················· | ···················· | ···················· |
| P | 7316 hCoV-19/England/ALDP-AA07B8/2020\|EPI_ISL_623996\|20 | ···················· | ···················· | ···················· | ···················· | ···················· | ···················· |
| P | 7317 hCoV-19/England/ALDP-AA33A2/2020\|EPI_ISL_623997\|20 | ···················· | ···················· | ···················· | ···················· | ···················· | ···················· |
| P | 7318 hCoV-19/England/ALDP-AA4101/2020\|EPI_ISL_623998\|20 | ···················· | ···················· | ···················· | ···················· | ···················· | ···················· |
| P | 7319 hCoV-19/England/ALDP-AA0FB0/2020\|EPI_ISL_624318\|20 | ···················· | ···················· | ···················· | ···················· | ···················· | ···················· |
| P | 7320 hCoV-19/England/ALDP-AA4374/2020\|EPI_ISL_624319\|20 | ···················· | ···················· | ···················· | ···················· | ···················· | ···················· |
| P | 7321 hCoV-19/England/ALDP-AA3DE0/2020\|EPI_ISL_624320\|20 | ···················· | ···················· | ···················· | ···················· | ···················· | ···················· |
| P | 7322 hCoV-19/England/ALDP-AA10F8/2020\|EPI_ISL_624321\|20 | ···················· | ···················· | ···················· | ···················· | ···················· | ···················· |
| P | 7323 hCoV-19/England/ALDP-AA0F65/2020\|EPI_ISL_624322\|20 | ···················· | ···················· | ···················· | ···················· | ···················· | ···················· |
| P | 7324 hCoV-19/England/ALDP-AA3D3B/2020\|EPI_ISL_624323\|20 | ···················· | ·A·················· | ···················· | ···················· | ···················· | ··A················· |
| P | 7325 hCoV-19/England/ALDP-AA217C/2020\|EPI_ISL_624324\|20 | ···················· | ···················· | ···················· | ···················· | ···················· | ···················· |
| P | 7326 hCoV-19/England/ALDP-AA2DB4/2020\|EPI_ISL_624325\|20 | ···················· | ·A·················· | ···················· | ···················· | ···················· | ··A················· |
| P | 7327 hCoV-19/England/ALDP-AA421D/2020\|EPI_ISL_624326\|20 | ···················· | ···················· | ···················· | ···················· | ···················· | ···················· |
| P | 7328 hCoV-19/England/ALDP-AA3ABC/2020\|EPI_ISL_624327\|20 | ···················· | ···················· | ···················· | ···················· | ···················· | ···················· |
| P | 7329 hCoV-19/England/ALDP-AA463F/2020\|EPI_ISL_624328\|20 | ···················· | ···················· | ···················· | ···················· | ···················· | ···················· |
| P | 7330 hCoV-19/England/ALDP-AA23B2/2020\|EPI_ISL_624330\|20 | ···················· | ···················· | ···················· | ···················· | ···················· | ···················· |
| P | 7331 hCoV-19/England/ALDP-AA47C3/2020\|EPI_ISL_624331\|20 | ···················· | ···················· | ···················· | ···················· | ···················· | ···················· |
| P | 7332 hCoV-19/England/ALDP-AA208E/2020\|EPI_ISL_624332\|20 | ···················· | ···················· | ···················· | ···················· | ···················· | ···················· |
| P | 7333 hCoV-19/England/ALDP-AA39BF/2020\|EPI_ISL_624333\|20 | ···················· | ···················· | ···················· | ···················· | ···················· | ···················· |
| P | 7334 hCoV-19/England/ALDP-AA3CB6/2020\|EPI_ISL_624334\|20 | ···················· | ···················· | ···················· | ···················· | ···················· | ···················· |
| P | 7335 hCoV-19/England/ALDP-AA1070/2020\|EPI_ISL_624336\|20 | ···················· | ···················· | ···················· | ···················· | ···················· | ···················· |
| P | 7336 hCoV-19/England/ALDP-AA4462/2020\|EPI_ISL_624337\|20 | ···················· | ···················· | ···················· | ···················· | ···················· | ···················· |
| P | 7337 hCoV-19/England/ALDP-AA39CE/2020\|EPI_ISL_624338\|20 | ···················· | ···················· | ···················· | ···················· | ···················· | ···················· |
| P | 7338 hCoV-19/England/ALDP-AA2051/2020\|EPI_ISL_624339\|20 | ···················· | ···················· | ···················· | ···················· | ···················· | ···················· |
| P | 7339 hCoV-19/England/ALDP-AA3DD1/2020\|EPI_ISL_624340\|20 | ···················· | ···················· | ···················· | ···················· | ···················· | ···················· |
| P | 7340 hCoV-19/England/ALDP-AA4B4F/2020\|EPI_ISL_624341\|20 | ···················· | ···················· | ···················· | ···················· | ···················· | ···················· |
| P | 7341 hCoV-19/England/ALDP-AA1034/2020\|EPI_ISL_624342\|20 | ···················· | ···················· | ···················· | ···················· | ···················· | ···················· |
| P | 7342 hCoV-19/England/ALDP-AA3B6E/2020\|EPI_ISL_624343\|20 | ···················· | ···················· | ···················· | ···················· | ···················· | ···················· |
| P | 7343 hCoV-19/England/ALDP-AA45C9/2020\|EPI_ISL_624344\|20 | ···················· | ···················· | ···················· | ···················· | ···················· | ···················· |
| P | 7344 hCoV-19/England/ALDP-AA1544/2020\|EPI_ISL_624345\|20 | ···················· | ···················· | ···················· | ···················· | ···················· | ···················· |
| P | 7345 hCoV-19/England/ALDP-AA472D/2020\|EPI_ISL_624346\|20 | ···················· | ···················· | ···················· | ···················· | ···················· | ···················· |
| P | 7346 hCoV-19/England/ALDP-AA4C00/2020\|EPI_ISL_624347\|20 | ···················· | ···················· | ···················· | ···················· | ···················· | ···················· |
| P | 7347 hCoV-19/England/ALDP-AA46B7/2020\|EPI_ISL_624348\|20 | ···················· | ···················· | ···················· | ···················· | ···················· | ···················· |
| P | 7348 hCoV-19/Wales/ALDP-AA2349/2020\|EPI_ISL_624349\|2020 | ···················· | ···················· | ···················· | ···················· | ···················· | ···················· |
| P | 7349 hCoV-19/England/ALDP-AA1ADC/2020\|EPI_ISL_624351\|20 | ···················· | ···················· | ···················· | ···················· | ···················· | ···················· |
| P | 7350 hCoV-19/England/ALDP-AA4602/2020\|EPI_ISL_624352\|20 | ···················· | ···················· | ···················· | ···················· | ···················· | ···················· |
| P | 7351 hCoV-19/England/ALDP-AA1F37/2020\|EPI_ISL_624353\|20 | ···················· | ···················· | ···················· | ···················· | ···················· | ···················· |
| P | 7352 hCoV-19/England/ALDP-AA22D3/2020\|EPI_ISL_624355\|20 | ···················· | ·A·················· | ···················· | ···················· | ···················· | ··A················· |
| P | 7353 hCoV-19/England/ALDP-AA3ACB/2020\|EPI_ISL_624357\|20 | ···················· | ···················· | ···················· | ···················· | ···················· | ···················· |
| P | 7354 hCoV-19/England/ALDP-AA468A/2020\|EPI_ISL_624358\|20 | ···················· | ···················· | ···················· | ···················· | ···················· | ···················· |
| P | 7355 hCoV-19/England/ALDP-AA459C/2020\|EPI_ISL_624359\|20 | ···················· | ···················· | ···················· | ···················· | ···················· | ···················· |
| P | 7356 hCoV-19/England/ALDP-AA2473/2020\|EPI_ISL_624360\|20 | ···················· | ···················· | ···················· | ···················· | ···················· | ···················· |
| P | 7357 hCoV-19/England/ALDP-AA2798/2020\|EPI_ISL_624361\|20 | ···················· | ···················· | ···················· | ···················· | ···················· | ···················· |
| P | 7358 hCoV-19/England/ALDP-AA4B6D/2020\|EPI_ISL_624362\|20 | ···················· | ···················· | ···················· | ···················· | ···················· | ···················· |
| P | 7359 hCoV-19/England/ALDP-AA1CC7/2020\|EPI_ISL_624363\|20 | ···················· | ···················· | ···················· | ···················· | ···················· | ···················· |
| P | 7360 hCoV-19/England/ALDP-AA4392/2020\|EPI_ISL_624364\|20 | ···················· | ···················· | ···················· | ···················· | ···················· | ···················· |
| P | 7361 hCoV-19/England/ALDP-AA3E0B/2020\|EPI_ISL_624365\|20 | ···················· | ···················· | ···················· | ···················· | ···················· | ···················· |
| P | 7362 hCoV-19/England/ALDP-AA27C5/2020\|EPI_ISL_624366\|20 | ···················· | ···················· | ···················· | ···················· | ···················· | ···················· |
| P | 7363 hCoV-19/England/ALDP-AA109E/2020\|EPI_ISL_624367\|20 | ···················· | ···················· | ···················· | ···················· | ···················· | ···················· |
| P | 7364 hCoV-19/England/ALDP-AA1A27/2020\|EPI_ISL_624368\|20 | ···················· | ···················· | ···················· | ···················· | ···················· | ···················· |
| P | 7365 hCoV-19/England/ALDP-AA1CA9/2020\|EPI_ISL_624369\|20 | ···················· | ···················· | ···················· | ···················· | ···················· | ···················· |
| P | 7366 hCoV-19/England/ALDP-AA3D0E/2020\|EPI_ISL_624370\|20 | ···················· | ···················· | ···················· | ···················· | ···················· | ···················· |

| | | | | | | | |
|---|---|---|---|---|---|---|---|
| P | 7367 hCoV-19/England/ALDP-AA2886/2020\|EPI_ISL_624371\|20 | ···················· | ···················· | ···················· | ···················· | ···················· | ···················· |
| P | 7368 hCoV-19/England/ALDP-AA15F9/2020\|EPI_ISL_624373\|20 | ···················· | ···················· | ···················· | ···················· | ···················· | ···················· |
| P | 7369 hCoV-19/England/ALDP-AA2428/2020\|EPI_ISL_624374\|20 | ···················· | ···················· | ···················· | ···················· | ···················· | ···················· |
| P | 7370 hCoV-19/England/ALDP-AA3A70/2020\|EPI_ISL_624375\|20 | ···················· | ···················· | ···················· | ···················· | ···················· | ···················· |
| P | 7371 hCoV-19/England/ALDP-AA0FFC/2020\|EPI_ISL_624376\|20 | ···················· | ···················· | ···················· | ···················· | ···················· | ···················· |
| P | 7372 hCoV-19/England/ALDP-AA471E/2020\|EPI_ISL_624377\|20 | ···················· | ···················· | ···················· | ···················· | ···················· | ···················· |
| P | 7373 hCoV-19/England/ALDP-AA276B/2020\|EPI_ISL_624378\|20 | ···················· | ···················· | ···················· | ···················· | ···················· | ···················· |
| P | 7374 hCoV-19/England/ALDP-AA1E94/2020\|EPI_ISL_624379\|20 | ···················· | ·A·················· | ···················· | ···················· | ···················· | ···A················ |
| P | 7375 hCoV-19/England/ALDP-AA42E0/2020\|EPI_ISL_624380\|20 | ···················· | ···················· | ···················· | ···················· | ···················· | ···················· |
| P | 7376 hCoV-19/England/ALDP-AA3D77/2020\|EPI_ISL_624381\|20 | ···T················ | ···················· | ···················· | ···················· | ···················· | ···················· |
| P | 7377 hCoV-19/England/ALDP-AA1EFE/2020\|EPI_ISL_624382\|20 | ···················· | ···················· | ···················· | ···················· | ······T············· | ···················· |
| P | 7378 hCoV-19/England/ALDP-AA27E3/2020\|EPI_ISL_624383\|20 | ···················· | ···················· | ···················· | ···················· | ···················· | ···················· |
| P | 7379 hCoV-19/England/ALDP-AA1ACD/2020\|EPI_ISL_624384\|20 | ···················· | ···················· | ···················· | ···················· | ···················· | ···················· |
| P | 7380 hCoV-19/England/ALDP-AA43B0/2020\|EPI_ISL_624385\|20 | ···················· | ···················· | ···················· | ···················· | ···················· | ···················· |
| P | 7381 hCoV-19/England/ALDP-AA4471/2020\|EPI_ISL_624386\|20 | ···················· | ···················· | ···················· | ···················· | ···················· | ···················· |
| P | 7382 hCoV-19/England/ALDP-AA39EC/2020\|EPI_ISL_624387\|20 | ···················· | ···················· | ···················· | ···················· | ···················· | ···················· |
| P | 7383 hCoV-19/England/ALDP-AA1F0A/2020\|EPI_ISL_624388\|20 | ···················· | ···················· | ···················· | ···················· | ···················· | ···················· |
| P | 7384 hCoV-19/England/ALDP-AA1A81/2020\|EPI_ISL_624389\|20 | ···················· | ···················· | ···················· | ···················· | ···················· | ···················· |
| P | 7385 hCoV-19/England/ALDP-AA2024/2020\|EPI_ISL_624390\|20 | ···················· | ···················· | ···················· | ···················· | ···················· | ···················· |
| P | 7386 hCoV-19/England/ALDP-AA2015/2020\|EPI_ISL_624391\|20 | ···················· | ···················· | ···················· | ···················· | ···················· | ···················· |
| P | 7387 hCoV-19/England/ALDP-AA2455/2020\|EPI_ISL_624392\|20 | ···················· | ···················· | ···················· | ···················· | ···················· | ···················· |
| P | 7388 hCoV-19/England/ALDP-AA1D88/2020\|EPI_ISL_624393\|20 | ···················· | ···················· | ···················· | ···················· | ···················· | ···················· |
| P | 7389 hCoV-19/England/ALDP-AA424A/2020\|EPI_ISL_624394\|20 | ···················· | ···················· | ···················· | ···················· | ···················· | ···················· |
| P | 7390 hCoV-19/England/ALDP-AA1FCE/2020\|EPI_ISL_624395\|20 | ···················· | ···················· | ···················· | ···················· | ···················· | ···················· |
| P | 7391 hCoV-19/England/ALDP-AA2491/2020\|EPI_ISL_624396\|20 | ···················· | ···················· | ···················· | ···················· | ···················· | ···················· |
| P | 7392 hCoV-19/Wales/ALDP-AA480C/2020\|EPI_ISL_624397\|2020 | ···················· | ···················· | ···················· | ···················· | ···················· | ···················· |
| P | 7393 hCoV-19/England/ALDP-AA2367/2020\|EPI_ISL_624398\|20 | ···················· | ···················· | ···················· | ···················· | ···················· | ···················· |
| P | 7394 hCoV-19/Wales/ALDP-AA4918/2020\|EPI_ISL_624399\|2020 | ···················· | ···················· | ···················· | ···················· | ···················· | ···················· |
| P | 7395 hCoV-19/USA/CA-CZB-11807/2020\|EPI_ISL_625649\|2020- | ···················· | ···················· | ···················· | ···················· | ···················· | ···················· |
| P | 7396 hCoV-19/USA/CA-CZB-11808/2020\|EPI_ISL_625650\|2020- | ···················· | ···················· | ···················· | ···················· | ···················· | ···················· |
| P | 7397 hCoV-19/USA/CA-CZB-11809/2020\|EPI_ISL_625651\|2020- | ···················· | ···················· | ···················· | ···················· | ···················· | ···················· |
| P | 7398 hCoV-19/USA/CA-CZB-11811/2020\|EPI_ISL_625652\|2020- | ···················· | ···················· | ···················· | ···················· | ···················· | ···················· |
| P | 7399 hCoV-19/USA/CA-CZB-11815/2020\|EPI_ISL_625654\|2020- | ···················· | ···················· | ···················· | ···················· | ···················· | ···················· |
| P | 7400 hCoV-19/USA/CA-CZB-11816/2020\|EPI_ISL_625655\|2020- | ···················· | ···················· | ···················· | ···················· | ···················· | ···················· |
| P | 7401 hCoV-19/USA/CA-CZB-11817/2020\|EPI_ISL_625656\|2020- | ···················· | ···················· | ···················· | ···················· | ···················· | ···················· |
| P | 7402 hCoV-19/USA/CA-CZB-11818/2020\|EPI_ISL_625657\|2020- | ···················· | ···················· | ···················· | ···················· | ···················· | ···················· |
| P | 7403 hCoV-19/USA/CA-CZB-11781/2020\|EPI_ISL_625660\|2020- | ···················· | ···················· | ···················· | ···················· | ···················· | ···················· |
| P | 7405 hCoV-19/USA/CA-CZB-11957/2020\|EPI_ISL_625663\|2020- | ···················· | ···················· | ···················· | ···················· | ···················· | ···················· |
| P | 7406 hCoV-19/USA/CA-CZB-11956/2020\|EPI_ISL_625664\|2020- | ···················· | ···················· | ···················· | ···················· | ···················· | ···················· |
| P | 7407 hCoV-19/USA/CA-CZB-11954/2020\|EPI_ISL_625665\|2020- | ···T················ | ·A·················· | ···················· | ···················· | ···················· | ···················· |
| P | 7408 hCoV-19/USA/CA-CZB-11953/2020\|EPI_ISL_625666\|2020- | ···················· | ·A·················· | ···················· | ···················· | ···················· | ···················· |
| P | 7409 hCoV-19/USA/CA-CZB-11950/2020\|EPI_ISL_625667\|2020- | ···················· | ···················· | ···················· | ···················· | ···················· | ···················· |
| P | 7410 hCoV-19/USA/CA-CZB-11949/2020\|EPI_ISL_625668\|2020- | ···T················ | ·A·················· | ···················· | ···················· | ···················· | ···················· |
| P | 7411 hCoV-19/USA/CA-CZB-11938/2020\|EPI_ISL_625669\|2020- | ···················· | ···················· | ···················· | ···················· | ···················· | ···················· |
| P | 7412 hCoV-19/USA/CA-CZB-11778/2020\|EPI_ISL_625670\|2020- | ···················· | ·············A······ | ···················· | ···················· | ···················· | ···················· |
| P | 7413 hCoV-19/Chile/MA-UMAG-Catg-5/2020\|EPI_ISL_625673\|2 | ···················· | ···················· | ···················· | ···················· | ···················· | ···················· |
| P | 7414 hCoV-19/Chile/MA-UMAG-Catg-6/2020\|EPI_ISL_625674\|2 | ···················· | ···················· | ···················· | ···················· | ···················· | ···················· |
| P | 7415 hCoV-19/Chile/MA-UMAG-Catg-7/2020\|EPI_ISL_625675\|2 | ···················· | ···················· | ···················· | ···················· | ···················· | ···················· |
| P | 7416 hCoV-19/Chile/MA-UMAG-Catg-20/2020\|EPI_ISL_625676\| | ···················· | ···················· | ···················· | ···················· | ···················· | ···················· |
| P | 7417 hCoV-19/Chile/MA-UMAG-Catg-21/2020\|EPI_ISL_625677\| | ···················· | ···················· | ···················· | ···················· | ···················· | ···················· |
| P | 7418 hCoV-19/Chile/MA-UMAG-Catg-4/2020\|EPI_ISL_625678\|2 | ···················· | ···················· | ···················· | ···················· | ···················· | ···················· |
| P | 7419 hCoV-19/Chile/MA-UMAG-Catg-5-2/2020\|EPI_ISL_625679 | ···················· | ···················· | ···················· | ···················· | ···················· | ···················· |
| P | 7420 hCoV-19/Chile/MA-UMAG-Catg-19/2020\|EPI_ISL_625680\| | ···················· | ···················· | ···················· | ···················· | ···················· | ···················· |
| P | 7421 hCoV-19/Chile/MA-UMAG-Catg-2/2020\|EPI_ISL_625681\|2 | ···················· | ···················· | ···················· | ···················· | ···················· | ···················· |
| P | 7422 hCoV-19/Chile/MA-UMAG-Catg-3/2020\|EPI_ISL_625682\|2 | ···················· | ···················· | ···················· | ···················· | ···················· | ···················· |
| P | 7423 hCoV-19/Denmark/DCGC-8554/2020\|EPI_ISL_625684\|2020 | ···················· | ·A·················· | ···················· | ···················· | ···················· | ···················· |
| P | 7424 hCoV-19/Denmark/DCGC-8644/2020\|EPI_ISL_625697\|2020 | ···················· | ···················· | ···················· | ···················· | ···················· | ···················· |
| P | 7425 hCoV-19/Switzerland/BS-ETHZ-330424/2020\|EPI_ISL_61 | ···················· | ···················· | ···················· | ···················· | ···················· | ···················· |
| P | 7426 hCoV-19/Switzerland/BS-ETHZ-330425/2020\|EPI_ISL_61 | ···················· | ···················· | ···················· | ···················· | ···················· | ···················· |
| P | 7427 hCoV-19/Switzerland/BE-ETHZ-330427/2020\|EPI_ISL_61 | ···················· | ···················· | ···················· | ···················· | ···················· | ···················· |
| P | 7428 hCoV-19/Switzerland/BE-ETHZ-330432/2020\|EPI_ISL_61 | ···················· | ···················· | ···················· | ···················· | ···················· | ···················· |
| P | 7429 hCoV-19/Switzerland/BE-ETHZ-330510/2020\|EPI_ISL_61 | ···················· | ···················· | ···················· | ···················· | ···················· | ···················· |
| P | 7430 hCoV-19/Denmark/DCGC-8036/2020\|EPI_ISL_617150\|2020 | ···················· | ···················· | ···················· | ···················· | ···················· | ···················· |
| P | 7431 hCoV-19/Denmark/DCGC-8042/2020\|EPI_ISL_617151\|2020 | ···················· | ···················· | ···················· | ···················· | ···················· | ···················· |
| P | 7432 hCoV-19/Denmark/DCGC-8064/2020\|EPI_ISL_617152\|2020 | ···················· | ·A·················· | ···················· | ···················· | ···················· | ···················· |
| P | 7433 hCoV-19/Denmark/DCGC-8069/2020\|EPI_ISL_617153\|2020 | ···················· | ···················· | ···················· | ···················· | ···················· | ···················· |
| P | 7434 hCoV-19/Denmark/DCGC-8077/2020\|EPI_ISL_617154\|2020 | ···················· | ···················· | ···················· | ···················· | ···················· | ···················· |
| P | 7435 hCoV-19/Denmark/DCGC-8083/2020\|EPI_ISL_617155\|2020 | ···················· | ···················· | ···················· | ···················· | ···················· | ···················· |
| P | 7436 hCoV-19/Denmark/DCGC-8093/2020\|EPI_ISL_617156\|2020 | ···················· | ·A·················· | ···················· | ···················· | ···················· | ···················· |
| P | 7437 hCoV-19/Denmark/DCGC-8096/2020\|EPI_ISL_617157\|2020 | ···················· | ···················· | ···················· | ···················· | ···················· | ···················· |
| P | 7438 hCoV-19/Denmark/DCGC-8043/2020\|EPI_ISL_617158\|2020 | ···················· | ···················· | ···················· | ···················· | ···················· | ···················· |
| P | 7439 hCoV-19/Denmark/DCGC-8050/2020\|EPI_ISL_617159\|2020 | ···················· | ···················· | ···················· | ···················· | ···················· | ···················· |
| P | 7440 hCoV-19/Denmark/DCGC-8058/2020\|EPI_ISL_617160\|2020 | ···················· | ···················· | ···················· | ···················· | ···················· | ···················· |
| P | 7441 hCoV-19/Denmark/DCGC-8065/2020\|EPI_ISL_617161\|2020 | ···················· | ·A·················· | ···················· | ···················· | ···················· | ···················· |
| P | 7442 hCoV-19/Denmark/DCGC-8084/2020\|EPI_ISL_617162\|2020 | ···················· | ···················· | ···················· | ···················· | ···················· | ···················· |
| P | 7443 hCoV-19/Denmark/DCGC-8102/2020\|EPI_ISL_617163\|2020 | ···················· | ·A·················· | ···················· | ···················· | ···················· | ···················· |
| P | 7444 hCoV-19/Denmark/DCGC-8051/2020\|EPI_ISL_617164\|2020 | ···················· | ···················· | ···················· | ···················· | ···················· | ···················· |
| P | 7445 hCoV-19/Denmark/DCGC-8071/2020\|EPI_ISL_617165\|2020 | ···················· | ···················· | ···················· | ···················· | ···················· | ···················· |
| P | 7446 hCoV-19/Denmark/DCGC-8085/2020\|EPI_ISL_617166\|2020 | ···················· | ···················· | ···················· | ···················· | ···················· | ···················· |
| P | 7447 hCoV-19/Denmark/DCGC-8103/2020\|EPI_ISL_617167\|2020 | ···················· | ·A·················· | ···················· | ···················· | ···················· | ···················· |

P 7448 hCoV-19/Denmark/DCGC-8108/2020|EPI_ISL_617168|2020 .................... .................... .................... .................... .................... ....................
P 7449 hCoV-19/Denmark/DCGC-8045/2020|EPI_ISL_617169|2020 .................... .................... .................... .................... .................... ....................
P 7450 hCoV-19/Denmark/DCGC-8052/2020|EPI_ISL_617170|2020 .................... .................... .................... .................... .................... ....................
P 7451 hCoV-19/Denmark/DCGC-8066/2020|EPI_ISL_617171|2020 .................... .A.................. .................... .................... .................... ....................
P 7452 hCoV-19/Denmark/DCGC-8072/2020|EPI_ISL_617172|2020 .................... .................... .................... .................... .................... ....................
P 7453 hCoV-19/Denmark/DCGC-8078/2020|EPI_ISL_617173|2020 .................... .................... .................... .................... .................... ....................
P 7454 hCoV-19/Denmark/DCGC-8053/2020|EPI_ISL_617174|2020 .................... .................... .................... .................... .................... ....................
P 7455 hCoV-19/Denmark/DCGC-8060/2020|EPI_ISL_617175|2020 .................... .................... .................... .................... .................... ....................
P 7456 hCoV-19/Denmark/DCGC-8079/2020|EPI_ISL_617176|2020 .................... .................... .................... .................... .................... ....................
P 7457 hCoV-19/Denmark/DCGC-8090/2020|EPI_ISL_617177|2020 .................... .................... .................... .................... .................... ....................
P 7458 hCoV-19/Denmark/DCGC-8099/2020|EPI_ISL_617178|2020 .................... .A.................. .................... .................... .................... ....................
P 7459 hCoV-19/Denmark/DCGC-8109/2020|EPI_ISL_617179|2020 .................... .................... .................... .................... .................... ....................
P 7460 hCoV-19/Denmark/DCGC-8055/2020|EPI_ISL_617185|2020 .................... .................... .................... .................... .................... ....................
P 7461 hCoV-19/Denmark/DCGC-8062/2020|EPI_ISL_617186|2020 .................... .................... .................... .................... .................... ....................
P 7462 hCoV-19/Denmark/DCGC-8075/2020|EPI_ISL_617187|2020 .................... .A.................. .................... .................... .................... ....................
P 7463 hCoV-19/Denmark/DCGC-8086/2020|EPI_ISL_617189|2020 .................... .................... .................... .................... .................... ....................
P 7464 hCoV-19/Denmark/DCGC-8131/2020|EPI_ISL_617200|2020 .................... .................... .................... .................... .................... ....................
P 7465 hCoV-19/Denmark/DCGC-8137/2020|EPI_ISL_617201|2020 .................... .A.................. .................... .................... .................... ....................
P 7466 hCoV-19/Denmark/DCGC-8153/2020|EPI_ISL_617202|2020 .................... .................... .................... .................... .................... ....................
P 7467 hCoV-19/Denmark/DCGC-8167/2020|EPI_ISL_617203|2020 .................... .................... .................... .................... .................... ....................
P 7468 hCoV-19/Denmark/DCGC-8179/2020|EPI_ISL_617204|2020 .................... .................... .................... .................... .................... ....................
P 7469 hCoV-19/Denmark/DCGC-8110/2020|EPI_ISL_617205|2020 .................... .................... .................... .................... .................... ....................
P 7470 hCoV-19/Denmark/DCGC-8126/2020|EPI_ISL_617206|2020 .................... .................... .................... .................... .................... ....................
P 7471 hCoV-19/Denmark/DCGC-8146/2020|EPI_ISL_617207|2020 .................... .................... .................... .................... .................... ....................
P 7472 hCoV-19/Denmark/DCGC-8154/2020|EPI_ISL_617208|2020 .................... .................... .................... .................... .................... ....................
P 7473 hCoV-19/Denmark/DCGC-8162/2020|EPI_ISL_617209|2020 .................... .................... .................... .................... .................... ....................
P 7474 hCoV-19/Denmark/DCGC-8168/2020|EPI_ISL_617210|2020 .................... .................... .................... .................... .................... ....................
P 7475 hCoV-19/Denmark/DCGC-8180/2020|EPI_ISL_617211|2020 .................... .A.................. .................... .................... .................... ....................
P 7476 hCoV-19/Denmark/DCGC-8111/2020|EPI_ISL_617212|2020 .................... .................... .................... .................... .................... ....................
P 7477 hCoV-19/Denmark/DCGC-8127/2020|EPI_ISL_617213|2020 .................... .................... .................... .................... .................... ....................
P 7478 hCoV-19/Denmark/DCGC-8139/2020|EPI_ISL_617214|2020 .................... .................... .................... .................... .................... ....................
P 7479 hCoV-19/Denmark/DCGC-8147/2020|EPI_ISL_617215|2020 .................... .................... .................... .................... .................... ....................
P 7480 hCoV-19/Denmark/DCGC-8155/2020|EPI_ISL_617216|2020 .................... .................... .................... .................... .................... ....................
P 7481 hCoV-19/Denmark/DCGC-8173/2020|EPI_ISL_617217|2020 .................... .................... .................... .................... .................... ....................
P 7482 hCoV-19/Denmark/DCGC-8181/2020|EPI_ISL_617218|2020 .................... .A.................. .................... .................... .................... ....................
P 7483 hCoV-19/Denmark/DCGC-8112/2020|EPI_ISL_617219|2020 .................... .................... .................... .................... .................... ....................
P 7484 hCoV-19/Denmark/DCGC-8120/2020|EPI_ISL_617220|2020 .................... .................... .................... .................... .................... ....................
P 7485 hCoV-19/Denmark/DCGC-8133/2020|EPI_ISL_617221|2020 .................... .................... .................... .................... .................... ....................
P 7486 hCoV-19/Denmark/DCGC-8140/2020|EPI_ISL_617222|2020 .................... .................... .................... .................... .................... ....................
P 7487 hCoV-19/Denmark/DCGC-8156/2020|EPI_ISL_617223|2020 .................... .................... .................... .................... .................... ....................
P 7488 hCoV-19/Denmark/DCGC-8169/2020|EPI_ISL_617224|2020 .................... .................... .................... .................... .................... ....................
P 7489 hCoV-19/Denmark/DCGC-8174/2020|EPI_ISL_617225|2020 .................... .................... .................... .................... .................... ....................
P 7490 hCoV-19/Denmark/DCGC-8182/2020|EPI_ISL_617226|2020 .................... .................... .................... .................... .................... ....................
P 7491 hCoV-19/Denmark/DCGC-8189/2020|EPI_ISL_617227|2020 .................... .................... .................... .................... .................... ....................
P 7492 hCoV-19/Denmark/DCGC-8113/2020|EPI_ISL_617228|2020 .................... .................... .................... .................... .................... ....................
P 7493 hCoV-19/Denmark/DCGC-8134/2020|EPI_ISL_617229|2020 .................... .................... .................... .................... .................... ....................
P 7494 hCoV-19/Denmark/DCGC-7532/2020|EPI_ISL_618480|2020 .................... .................... .................... .................... .................... ....................
P 7495 hCoV-19/Denmark/DCGC-7544/2020|EPI_ISL_618481|2020 .................... .................... .................... .................... .................... ....................
P 7496 hCoV-19/Denmark/DCGC-7550/2020|EPI_ISL_618482|2020 .................... .................... .................... .................... .................... ....................
P 7497 hCoV-19/Denmark/DCGC-7575/2020|EPI_ISL_618485|2020 .................... .................... .................... .................... .................... ....................
P 7498 hCoV-19/Denmark/DCGC-7588/2020|EPI_ISL_618487|2020 .................... .................... .................... .................... .................... ....................
P 7499 hCoV-19/Denmark/DCGC-7533/2020|EPI_ISL_618489|2020 .................... .................... .................... .................... .................... ....................
P 7500 hCoV-19/Denmark/DCGC-7545/2020|EPI_ISL_618490|2020 .................... .................... .................... .................... .................... ....................
P 7501 hCoV-19/Denmark/DCGC-7551/2020|EPI_ISL_618491|2020 .................... .................... .................... .................... .................... ....................
P 7502 hCoV-19/Denmark/DCGC-7564/2020|EPI_ISL_618492|2020 .................... .................... .................... .................... .................... ....................
P 7503 hCoV-19/Denmark/DCGC-7570/2020|EPI_ISL_618493|2020 .................... .................... .................... .................... .................... ....................
P 7504 hCoV-19/Denmark/DCGC-7582/2020|EPI_ISL_618495|2020 .................... .................... .................... .................... .................... ....................
P 7505 hCoV-19/Denmark/DCGC-1606/2020|EPI_ISL_618496|2020 .................... .................... .................... .................... .................... ....................
P 7506 hCoV-19/Denmark/DCGC-7589/2020|EPI_ISL_618497|2020 .................... .A.................. .................... .................... .................... ....................
P 7507 hCoV-19/Denmark/DCGC-7594/2020|EPI_ISL_618498|2020 .................... .................... .................... .................... .................... ....................
P 7508 hCoV-19/Denmark/DCGC-7599/2020|EPI_ISL_618499|2020 .................... .................... .................... .................... .................... ....................
P 7509 hCoV-19/Denmark/DCGC-7604/2020|EPI_ISL_618500|2020 .................... .................... .................... .................... .................... ....................
P 7510 hCoV-19/Denmark/DCGC-7611/2020|EPI_ISL_618501|2020 .................... .................... .................... .................... .................... ....................
P 7511 hCoV-19/Denmark/DCGC-7619/2020|EPI_ISL_618502|2020 .................... .................... .................... .................... .................... ....................
P 7512 hCoV-19/Denmark/DCGC-7624/2020|EPI_ISL_618503|2020 .................... .................... .................... .................... .................... ....................
P 7513 hCoV-19/Denmark/DCGC-7634/2020|EPI_ISL_618505|2020 .................... .................... .................... .................... .................... ....................
P 7514 hCoV-19/Denmark/DCGC-7651/2020|EPI_ISL_618507|2020 .................... ...A................ .................... .................... .................... ....................
P 7515 hCoV-19/Denmark/DCGC-7600/2020|EPI_ISL_618508|2020 .................... .................... .................... .................... .................... ....................
P 7516 hCoV-19/Denmark/DCGC-7615/2020|EPI_ISL_618510|2020 .................... .................... .................... .................... .................... ....................
P 7517 hCoV-19/Denmark/DCGC-1557/2020|EPI_ISL_618511|2020 .................... .................... .................... .................... .................... ....................
P 7518 hCoV-19/Denmark/DCGC-7641/2020|EPI_ISL_618513|2020 .................... .................... .................... .................... .................... ....................
P 7519 hCoV-19/Denmark/DCGC-7644/2020|EPI_ISL_618514|2020 .................... .................... .................... .................... .................... ....................
P 7520 hCoV-19/Denmark/DCGC-7652/2020|EPI_ISL_618515|2020 .................... .................... .................... .................... .................... ....................
P 7521 hCoV-19/Denmark/DCGC-7601/2020|EPI_ISL_618516|2020 .................... .................... .................... .................... .................... ....................
P 7522 hCoV-19/Denmark/DCGC-1558/2020|EPI_ISL_618517|2020 .................... .................... .................... .................... .................... ....................
P 7523 hCoV-19/Denmark/DCGC-7625/2020|EPI_ISL_618518|2020 .................... .................... .................... .................... .................... ....................
P 7524 hCoV-19/Denmark/DCGC-7646/2020|EPI_ISL_618530|2020 .................... .................... .................... .................... .................... ....................
P 7525 hCoV-19/Denmark/DCGC-7654/2020|EPI_ISL_618531|2020 .................... .................... .................... .................... .................... ....................
P 7526 hCoV-19/Denmark/DCGC-7602/2020|EPI_ISL_618532|2020 .................... .................... .................... .................... .................... ....................
P 7527 hCoV-19/Denmark/DCGC-7613/2020|EPI_ISL_618534|2020 .................... .................... .................... .................... .................... ....................

P 7528 hCoV-19/Denmark/DCGC-7621/2020|EPI_ISL_618536|2020 ···················· ···················· ····················· ···················· ···················· ····················
P 7529 hCoV-19/Denmark/DCGC-7627/2020|EPI_ISL_618537|2020 ···················· ···················· ····················· ···················· ···················· ····················
P 7530 hCoV-19/Denmark/DCGC-7638/2020|EPI_ISL_618538|2020 ···················· ···················· ····················· ···················· ···················· ····················
P 7531 hCoV-19/Denmark/DCGC-7647/2020|EPI_ISL_618539|2020 ···················· ···················· ····················· ···················· ···················· ····················
P 7532 hCoV-19/Denmark/DCGC-7648/2020|EPI_ISL_618540|2020 ···················· ·A·················· ····················· ···················· ···················· ····················
P 7533 hCoV-19/Denmark/DCGC-7639/2020|EPI_ISL_618544|2020 ···················· ···················· ····················· ···················· ···················· ····················
P 7534 hCoV-19/Denmark/DCGC-7649/2020|EPI_ISL_618545|2020 ···················· ·A·················· ····················· ···················· ···················· ····················
P 7535 hCoV-19/Denmark/DCGC-7617/2020|EPI_ISL_618547|2020 ···················· ···················· ····················· ···················· ···················· ····················
P 7536 hCoV-19/Denmark/DCGC-7622/2020|EPI_ISL_618548|2020 ···················· ···················· ····················· ···················· ···················· ····················
P 7537 hCoV-19/Denmark/DCGC-7629/2020|EPI_ISL_618549|2020 ···················· ···················· ····················· ···················· ···················· ····················
P 7538 hCoV-19/Denmark/DCGC-7603/2020|EPI_ISL_618550|2020 ···················· ···················· ····················· ···················· ···················· ····················
P 7539 hCoV-19/Denmark/DCGC-7618/2020|EPI_ISL_618552|2020 ···················· ···················· ····················· ···················· ···················· ····················
P 7540 hCoV-19/Denmark/DCGC-7623/2020|EPI_ISL_618553|2020 ···················· ···················· ····················· ···················· ···················· ····················
P 7541 hCoV-19/Denmark/DCGC-7633/2020|EPI_ISL_618555|2020 ···················· ···················· ····················· ···················· ···················· ····················
P 7542 hCoV-19/Denmark/DCGC-7640/2020|EPI_ISL_618556|2020 ···················· ···················· ····················· ···················· ···················· ····················
P 7543 hCoV-19/Denmark/DCGC-7650/2020|EPI_ISL_618557|2020 ···················· ···················· ····················· ···················· ···················· ····················
P 7544 hCoV-19/Denmark/DCGC-7655/2020|EPI_ISL_618558|2020 ···················· ···················· ····················· ···················· ···················· ····················
P 7545 hCoV-19/Denmark/DCGC-7663/2020|EPI_ISL_618559|2020 ···················· ···················· ····················· ···················· ···················· ····················
P 7546 hCoV-19/Denmark/DCGC-6962/2020|EPI_ISL_619808|2020 ···················· ···················· ····················· ···················· ···················· ····················
P 7547 hCoV-19/Denmark/DCGC-6967/2020|EPI_ISL_619813|2020 ···················· ···················· ····················· ···················· ···················· ····················
P 7548 hCoV-19/Denmark/DCGC-6968/2020|EPI_ISL_619814|2020 ···················· ···················· ····················· ···················· ···················· ····················
P 7549 hCoV-19/Denmark/DCGC-6970/2020|EPI_ISL_619816|2020 ···················· ···················· ····················· ···················· ···················· ····················
P 7550 hCoV-19/Denmark/DCGC-6971/2020|EPI_ISL_619817|2020 ···················· ···················· ····················· ···················· ···················· ····················
P 7551 hCoV-19/Denmark/DCGC-6972/2020|EPI_ISL_619818|2020 ···················· ·A·················· ····················· ···················· ···················· ····················
P 7552 hCoV-19/Denmark/DCGC-6973/2020|EPI_ISL_619819|2020 ···················· ···················· ····················· ···················· ···················· ····················
P 7553 hCoV-19/Denmark/DCGC-6974/2020|EPI_ISL_619820|2020 ···················· ···················· ····················· ···················· ···················· ····················
P 7554 hCoV-19/Denmark/DCGC-6975/2020|EPI_ISL_619821|2020 ···················· ···················· ····················· ···················· ···················· ····················
P 7555 hCoV-19/Denmark/DCGC-6976/2020|EPI_ISL_619822|2020 ···················· ···················· ····················· ···················· ···················· ····················
P 7556 hCoV-19/Denmark/DCGC-6977/2020|EPI_ISL_619823|2020 ···················· ···················· ····················· ···················· ···················· ····················
P 7557 hCoV-19/Denmark/DCGC-6978/2020|EPI_ISL_619824|2020 ···················· ···················· ····················· ···················· ···················· ····················
P 7558 hCoV-19/Denmark/DCGC-6979/2020|EPI_ISL_619825|2020 ···················· ···················· ····················· ···················· ···················· ····················
P 7559 hCoV-19/Denmark/DCGC-6980/2020|EPI_ISL_619826|2020 ···················· ···················· ····················· ···················· ···················· ····················
P 7560 hCoV-19/Denmark/DCGC-6981/2020|EPI_ISL_619827|2020 ···················· ···················· ····················· ···················· ···················· ····················
P 7561 hCoV-19/Denmark/DCGC-6982/2020|EPI_ISL_619828|2020 ···················· ···················· ····················· ···················· ···················· ····················
P 7562 hCoV-19/Denmark/DCGC-6984/2020|EPI_ISL_619830|2020 ···················· ···················· ····················· ···················· ···················· ····················
P 7563 hCoV-19/Denmark/DCGC-6985/2020|EPI_ISL_619831|2020 ···················· ···················· ····················· ···················· ···················· ····················
P 7564 hCoV-19/Denmark/DCGC-6987/2020|EPI_ISL_619833|2020 ···················· ···················· ····················· ···················· ···················· ····················
P 7565 hCoV-19/Denmark/DCGC-6988/2020|EPI_ISL_619834|2020 ···················· ···················· ····················· ···················· ···················· ····················
P 7566 hCoV-19/Denmark/DCGC-6989/2020|EPI_ISL_619835|2020 ···················· ···················· ····················· ···················· ···················· ····················
P 7567 hCoV-19/Denmark/DCGC-6990/2020|EPI_ISL_619836|2020 ···················· ···················· ····················· ···················· ···················· ····················
P 7568 hCoV-19/Denmark/DCGC-6991/2020|EPI_ISL_619837|2020 ···················· ···················· ····················· ···················· ···················· ····················
P 7569 hCoV-19/Denmark/DCGC-6993/2020|EPI_ISL_619839|2020 ···················· ···················· ····················· ···················· ···················· ····················
P 7570 hCoV-19/Denmark/DCGC-1566/2020|EPI_ISL_619840|2020 ···················· ···················· ····················· ···················· ···················· ····················
P 7571 hCoV-19/Denmark/DCGC-6995/2020|EPI_ISL_619842|2020 ···················· ···················· ····················· ···················· ···················· ····················
P 7572 hCoV-19/Denmark/DCGC-6996/2020|EPI_ISL_619843|2020 ···················· ···················· ····················· ···················· ···················· ····················
P 7573 hCoV-19/Denmark/DCGC-6997/2020|EPI_ISL_619844|2020 ···················· ···················· ····················· ···················· ···················· ····················
P 7574 hCoV-19/Denmark/DCGC-6998/2020|EPI_ISL_619845|2020 ···················· ···················· ····················· ···················· ···················· ····················
P 7575 hCoV-19/Denmark/DCGC-6999/2020|EPI_ISL_619846|2020 ···················· ···················· ····················· ···················· ···················· ····················
P 7576 hCoV-19/Denmark/DCGC-7000/2020|EPI_ISL_619847|2020 ···················· ···················· ····················· ···················· ···················· ····················
P 7577 hCoV-19/Denmark/DCGC-7001/2020|EPI_ISL_619848|2020 ···················· ···················· ····················· ···················· ···················· ····················
P 7578 hCoV-19/Denmark/DCGC-7002/2020|EPI_ISL_619849|2020 ···················· ·A·················· ····················· ···················· ···················· ··A·················
P 7579 hCoV-19/Denmark/DCGC-7003/2020|EPI_ISL_619850|2020 ···················· ···················· ····················· ···················· ···················· ····················
P 7580 hCoV-19/Denmark/DCGC-7013/2020|EPI_ISL_619860|2020 ···················· ···················· ····················· ···················· ···················· ····················
P 7581 hCoV-19/Denmark/DCGC-7014/2020|EPI_ISL_619861|2020 ···················· ···················· ····················· ···················· ···················· ····················
P 7582 hCoV-19/Denmark/DCGC-7015/2020|EPI_ISL_619862|2020 ···················· ···················· ····················· ···················· ···················· ····················
P 7583 hCoV-19/Denmark/DCGC-7017/2020|EPI_ISL_619864|2020 ···················· ···················· ····················· ···················· ···················· ····················
P 7584 hCoV-19/Denmark/DCGC-7018/2020|EPI_ISL_619865|2020 ···················· ···················· ····················· ···················· ···················· ····················
P 7585 hCoV-19/Denmark/DCGC-7019/2020|EPI_ISL_619866|2020 ···················· ···················· ····················· ···················· ···················· ····················
P 7586 hCoV-19/Denmark/DCGC-7021/2020|EPI_ISL_619868|2020 ···················· ···················· ····················· ···················· ···················· ····················
P 7587 hCoV-19/Denmark/DCGC-7022/2020|EPI_ISL_619869|2020 ···················· ········A··········· ····················· ···················· ···················· ····················
P 7588 hCoV-19/Denmark/DCGC-7023/2020|EPI_ISL_619870|2020 ···················· ···················· ····················· ···················· ···················· ····················
P 7589 hCoV-19/Denmark/DCGC-7024/2020|EPI_ISL_619871|2020 ···················· ·A·················· ····················· ···················· ···················· ··A·················
P 7590 hCoV-19/Denmark/DCGC-7025/2020|EPI_ISL_619872|2020 ···················· ···················· ····················· ···················· ···················· ····················
P 7591 hCoV-19/Denmark/DCGC-7026/2020|EPI_ISL_619873|2020 ···················· ···················· ····················· ···················· ···················· ····················
P 7592 hCoV-19/Denmark/DCGC-7030/2020|EPI_ISL_619877|2020 ···················· ···················· ····················· ···················· ···················· ····················
P 7593 hCoV-19/Denmark/DCGC-7032/2020|EPI_ISL_619879|2020 ···················· ···················· ····················· ···················· ···················· ····················
P 7594 hCoV-19/Denmark/DCGC-7033/2020|EPI_ISL_619880|2020 ···················· ···················· ····················· ···················· ···················· ····················
P 7595 hCoV-19/Denmark/DCGC-7034/2020|EPI_ISL_619881|2020 ···················· ···················· ····················· ···················· ···················· ····················
P 7596 hCoV-19/Denmark/DCGC-7035/2020|EPI_ISL_619882|2020 ···················· ···················· ····················· ···················· ···················· ····················
P 7597 hCoV-19/Denmark/DCGC-1645/2020|EPI_ISL_619885|2020 ···················· ···················· ····················· ···················· ···················· ····················
P 7598 hCoV-19/Denmark/DCGC-7038/2020|EPI_ISL_619886|2020 ···················· ···················· ····················· ···················· ···················· ····················
P 7599 hCoV-19/Denmark/DCGC-7039/2020|EPI_ISL_619887|2020 ···················· ···················· ····················· ···················· ···················· ····················
P 7600 hCoV-19/Denmark/DCGC-7041/2020|EPI_ISL_619889|2020 ···················· ···················· ····················· ···················· ···················· ····················
P 7601 hCoV-19/Denmark/DCGC-5475/2020|EPI_ISL_620800|2020 ···················· ···················· ····················· ···················· ···················· ····················
P 7602 hCoV-19/Denmark/DCGC-5476/2020|EPI_ISL_620801|2020 ···················· ···················· ····················· ···················· ······T············· ····················
P 7603 hCoV-19/Denmark/DCGC-5477/2020|EPI_ISL_620802|2020 ···················· ···················· ····················· ···················· ···················· ····················
P 7604 hCoV-19/Denmark/DCGC-5478/2020|EPI_ISL_620803|2020 ···················· ···················· ····················· ···················· ···················· ····················
P 7605 hCoV-19/Denmark/DCGC-5484/2020|EPI_ISL_620809|2020 ···················· ···················· ····················· ···················· ···················· ····················
P 7606 hCoV-19/Denmark/DCGC-5485/2020|EPI_ISL_620810|2020 ···················· ···················· ····················· ···················· ···················· ····················
P 7607 hCoV-19/Denmark/DCGC-5486/2020|EPI_ISL_620811|2020 ···················· ···················· ····················· ···················· ···················· ····················

| | | | | | | | | |
|---|---|---|---|---|---|---|---|---|
| P | 7608 | hCoV-19/Denmark/DCGC-5487/2020\|EPI_ISL_620812\|2020 | ···················· | ···················· | ···················· | ···················· | ···················· | ···················· |
| P | 7609 | hCoV-19/Denmark/DCGC-5488/2020\|EPI_ISL_620813\|2020 | ···················· | ···················· | ···················· | ···················· | ···················· | ···················· |
| P | 7610 | hCoV-19/Denmark/DCGC-5489/2020\|EPI_ISL_620814\|2020 | ···················· | ···················· | ···················· | ···················· | ···················· | ···················· |
| P | 7611 | hCoV-19/Denmark/DCGC-5490/2020\|EPI_ISL_620815\|2020 | ···················· | ···················· | ···················· | ···················· | ···················· | ···················· |
| P | 7612 | hCoV-19/Denmark/DCGC-5491/2020\|EPI_ISL_620816\|2020 | ···················· | ···················· | ···················· | ···················· | ···················· | ···················· |
| P | 7613 | hCoV-19/Denmark/DCGC-5492/2020\|EPI_ISL_620817\|2020 | ···················· | ···················· | ···················· | ···················· | ···················· | ···················· |
| P | 7614 | hCoV-19/Denmark/DCGC-5493/2020\|EPI_ISL_620818\|2020 | ···················· | ···················· | ···················· | ···················· | ···················· | ···················· |
| P | 7615 | hCoV-19/Denmark/DCGC-5494/2020\|EPI_ISL_620819\|2020 | ···················· | ···················· | ···················· | ···················· | ···················· | ···················· |
| P | 7616 | hCoV-19/Denmark/DCGC-5495/2020\|EPI_ISL_620820\|2020 | ···················· | ···················· | ···················· | ···················· | ···················· | ···················· |
| P | 7617 | hCoV-19/Denmark/DCGC-5496/2020\|EPI_ISL_620821\|2020 | ···················· | ···················· | ···················· | ···················· | ···················· | ···················· |
| P | 7618 | hCoV-19/Denmark/DCGC-5497/2020\|EPI_ISL_620822\|2020 | ···················· | ···················· | ···················· | ···················· | ···················· | ···················· |
| P | 7619 | hCoV-19/Denmark/DCGC-5498/2020\|EPI_ISL_620823\|2020 | ···················· | ···················· | ···················· | ···················· | ···················· | ···················· |
| P | 7620 | hCoV-19/Denmark/DCGC-5499/2020\|EPI_ISL_620824\|2020 | ···················· | ·A·················· | ···················· | ···················· | ···················· | ···················· |
| P | 7621 | hCoV-19/Denmark/DCGC-5500/2020\|EPI_ISL_620825\|2020 | ···················· | ···················· | ···················· | ···················· | ···················· | ···················· |
| P | 7622 | hCoV-19/Denmark/DCGC-5501/2020\|EPI_ISL_620826\|2020 | ···················· | ···················· | ···················· | ···················· | ···················· | ···················· |
| P | 7623 | hCoV-19/Denmark/DCGC-5502/2020\|EPI_ISL_620827\|2020 | ···················· | ···················· | ···················· | ···················· | ···················· | ···················· |
| P | 7624 | hCoV-19/Denmark/DCGC-5503/2020\|EPI_ISL_620828\|2020 | ···················· | ···················· | ···················· | ···················· | ···················· | ···················· |
| P | 7625 | hCoV-19/Denmark/DCGC-5504/2020\|EPI_ISL_620829\|2020 | ···················· | ···················· | ···················· | ···················· | ···················· | ···················· |
| P | 7626 | hCoV-19/Denmark/DCGC-5505/2020\|EPI_ISL_620830\|2020 | ···················· | ···················· | ···················· | ···················· | ···················· | ···················· |
| P | 7627 | hCoV-19/Denmark/DCGC-5506/2020\|EPI_ISL_620831\|2020 | ···················· | ···················· | ···················· | ···················· | ···················· | ···················· |
| P | 7628 | hCoV-19/Denmark/DCGC-5507/2020\|EPI_ISL_620832\|2020 | ···················· | ···················· | ···················· | ···················· | ···················· | ···················· |
| P | 7629 | hCoV-19/Denmark/DCGC-5508/2020\|EPI_ISL_620833\|2020 | ···················· | ···················· | ···················· | ···················· | ···················· | ···················· |
| P | 7630 | hCoV-19/Denmark/DCGC-5509/2020\|EPI_ISL_620834\|2020 | ···················· | ···················· | ···················· | ···················· | ···················· | ···················· |
| P | 7631 | hCoV-19/Denmark/DCGC-5510/2020\|EPI_ISL_620835\|2020 | ···················· | ···················· | ···················· | ···················· | ···················· | ···················· |
| P | 7632 | hCoV-19/Denmark/DCGC-5511/2020\|EPI_ISL_620836\|2020 | ···················· | ···················· | ···················· | ···················· | ···················· | ···················· |
| P | 7633 | hCoV-19/Denmark/DCGC-5512/2020\|EPI_ISL_620837\|2020 | ···················· | ···················· | ···················· | ···················· | ···················· | ···················· |
| P | 7634 | hCoV-19/Denmark/DCGC-5513/2020\|EPI_ISL_620838\|2020 | ···················· | ···················· | ···················· | ···················· | ···················· | ···················· |
| P | 7635 | hCoV-19/Denmark/DCGC-5514/2020\|EPI_ISL_620839\|2020 | ···················· | ···················· | ···················· | ···················· | ···················· | ···················· |
| P | 7636 | hCoV-19/Denmark/DCGC-5515/2020\|EPI_ISL_620840\|2020 | ···················· | ···················· | ···················· | ···················· | ···················· | ···················· |
| P | 7637 | hCoV-19/Denmark/DCGC-5516/2020\|EPI_ISL_620841\|2020 | ···················· | ···················· | ···················· | ···················· | ···················· | ···················· |
| P | 7638 | hCoV-19/Denmark/DCGC-5525/2020\|EPI_ISL_620850\|2020 | ···················· | ···················· | ···················· | ···················· | ···················· | ···················· |
| P | 7639 | hCoV-19/Denmark/DCGC-5526/2020\|EPI_ISL_620851\|2020 | ···················· | ···················· | ···················· | ···················· | ···················· | ···················· |
| P | 7640 | hCoV-19/Denmark/DCGC-5527/2020\|EPI_ISL_620852\|2020 | ···················· | ···················· | ···················· | ···················· | ···················· | ···················· |
| P | 7641 | hCoV-19/Denmark/DCGC-5528/2020\|EPI_ISL_620853\|2020 | ···················· | ···················· | ···················· | ···················· | ···················· | ···················· |
| P | 7642 | hCoV-19/Denmark/DCGC-5529/2020\|EPI_ISL_620854\|2020 | ···················· | ···················· | ···················· | ···················· | ···················· | ···················· |
| P | 7643 | hCoV-19/Denmark/DCGC-5530/2020\|EPI_ISL_620855\|2020 | ···················· | ···················· | ···················· | ···················· | ···················· | ···················· |
| P | 7644 | hCoV-19/Denmark/DCGC-5531/2020\|EPI_ISL_620856\|2020 | ···················· | ···················· | ···················· | ···················· | ···················· | ···················· |
| P | 7645 | hCoV-19/Denmark/DCGC-5532/2020\|EPI_ISL_620857\|2020 | ···················· | ···················· | ···················· | ···················· | ···················· | ···················· |
| P | 7646 | hCoV-19/Denmark/DCGC-5533/2020\|EPI_ISL_620858\|2020 | ···················· | ···················· | ···················· | ···················· | ···················· | ···················· |
| P | 7647 | hCoV-19/Denmark/DCGC-5534/2020\|EPI_ISL_620859\|2020 | ···················· | ···················· | ···················· | ···················· | ···················· | ···················· |
| P | 7648 | hCoV-19/Denmark/DCGC-5535/2020\|EPI_ISL_620860\|2020 | ···················· | ···················· | ···················· | ···················· | ···················· | ···················· |
| P | 7649 | hCoV-19/Denmark/DCGC-5536/2020\|EPI_ISL_620861\|2020 | ···················· | ···················· | ···················· | ···················· | ···················· | ···················· |
| P | 7650 | hCoV-19/Denmark/DCGC-5537/2020\|EPI_ISL_620862\|2020 | ···················· | ···················· | ···················· | ···················· | ···················· | ···················· |
| P | 7651 | hCoV-19/Denmark/DCGC-5538/2020\|EPI_ISL_620863\|2020 | ···················· | ···················· | ···················· | ···················· | ···················· | ···················· |
| P | 7652 | hCoV-19/Denmark/DCGC-5539/2020\|EPI_ISL_620864\|2020 | ···················· | ···················· | ···················· | ···················· | ···················· | ···················· |
| P | 7653 | hCoV-19/Denmark/DCGC-5540/2020\|EPI_ISL_620865\|2020 | ···················· | ···················· | ···················· | ···················· | ···················· | ···················· |
| P | 7654 | hCoV-19/Denmark/DCGC-5541/2020\|EPI_ISL_620866\|2020 | ···················· | ···················· | ···················· | ···················· | ···················· | ···················· |
| P | 7655 | hCoV-19/Denmark/DCGC-5542/2020\|EPI_ISL_620867\|2020 | ···················· | ···················· | ···················· | ···················· | ···················· | ···················· |
| P | 7656 | hCoV-19/Denmark/DCGC-5543/2020\|EPI_ISL_620868\|2020 | ···················· | ···················· | ···················· | ···················· | ···················· | ···················· |
| P | 7657 | hCoV-19/Denmark/DCGC-5544/2020\|EPI_ISL_620869\|2020 | ···················· | ···················· | ···················· | ···················· | ···················· | ···················· |
| P | 7658 | hCoV-19/South Africa/L00168/2020\|EPI_ISL_623073\|20 | ···················· | ···················· | ···················· | ···················· | ···················· | ···················· |
| P | 7659 | hCoV-19/Sweden/20-21502/2020\|EPI_ISL_623076\|2020-0 | ···················· | ···················· | ···················· | ···················· | ···················· | ···················· |
| P | 7660 | hCoV-19/Sweden/20-21503/2020\|EPI_ISL_623077\|2020-0 | ···················· | ···················· | ···················· | ···················· | ···················· | ···················· |
| P | 7661 | hCoV-19/Sweden/20-21504/2020\|EPI_ISL_623078\|2020-0 | ···················· | ···················· | ···················· | ···················· | ···················· | ···················· |
| P | 7662 | hCoV-19/Sweden/20-21505/2020\|EPI_ISL_623079\|2020-0 | ···················· | ···················· | ···················· | ···················· | ···················· | ···················· |
| P | 7663 | hCoV-19/Sweden/20-22147/2020\|EPI_ISL_623080\|2020-0 | ···················· | ···················· | ···················· | ···················· | ···················· | ···················· |
| P | 7664 | hCoV-19/Sweden/20-22148/2020\|EPI_ISL_623081\|2020-0 | ···················· | ···················· | ···················· | ···················· | ···················· | ···················· |
| P | 7665 | hCoV-19/Sweden/20-22149/2020\|EPI_ISL_623082\|2020-0 | ···················· | ···················· | ···················· | ···················· | ···················· | ···················· |
| P | 7666 | hCoV-19/Sweden/20-22152/2020\|EPI_ISL_623083\|2020-0 | ···················· | ···················· | ···················· | ···················· | ···················· | ···················· |
| P | 7667 | hCoV-19/Sweden/20-22153/2020\|EPI_ISL_623084\|2020-0 | ···················· | ···················· | ···················· | ···················· | ···················· | ···················· |
| P | 7668 | hCoV-19/Sweden/20-22156/2020\|EPI_ISL_623085\|2020-0 | ···················· | ···················· | ···················· | ···················· | ···················· | ···················· |
| P | 7669 | hCoV-19/Sweden/20-52028/2020\|EPI_ISL_623086\|2020-0 | ···················· | ···················· | ···················· | ···················· | ···················· | ···················· |
| P | 7670 | hCoV-19/Sweden/20-52054/2020\|EPI_ISL_623088\|2020-0 | ···················· | ···················· | ···················· | ···················· | ···················· | ···················· |
| P | 7671 | hCoV-19/Sweden/20-52075/2020\|EPI_ISL_623089\|2020-0 | ···················· | ···················· | ···················· | ···················· | ···················· | ···················· |
| P | 7672 | hCoV-19/Sweden/20-52117/2020\|EPI_ISL_623090\|2020-0 | ···················· | ···················· | ···················· | ···················· | ···················· | ···················· |
| P | 7673 | hCoV-19/Sweden/20-52162/2020\|EPI_ISL_623091\|2020-0 | ···················· | ···················· | ···················· | ···················· | ···················· | ···················· |
| P | 7674 | hCoV-19/Sweden/20-52703/2020\|EPI_ISL_623092\|2020-0 | ···················· | ···················· | ···················· | ···················· | ···················· | ···················· |
| P | 7675 | hCoV-19/Sweden/20-52704/2020\|EPI_ISL_623093\|2020-0 | ···················· | ···················· | ···················· | ···················· | ···················· | ···················· |
| P | 7676 | hCoV-19/Sweden/20-52707/2020\|EPI_ISL_623094\|2020-0 | ···················· | ···················· | ···················· | ···················· | ···················· | ···················· |
| P | 7677 | hCoV-19/Sweden/20-52801/2020\|EPI_ISL_623095\|2020-0 | ···················· | ···················· | ···················· | ···················· | ···················· | ···················· |
| P | 7678 | hCoV-19/France/BRE-9495/2020\|EPI_ISL_623096\|2020-1 | ···················· | ···················· | ···················· | ···················· | ···················· | ···················· |
| P | 7679 | hCoV-19/France/GES-9483/2020\|EPI_ISL_623097\|2020-1 | ···················· | ···················· | ···················· | ···················· | ···················· | ···················· |
| P | 7680 | hCoV-19/France/ARA-CDP24988/2020\|EPI_ISL_623099\|20 | ···················· | ···················· | ···················· | ···················· | ···················· | ···················· |
| P | 7681 | hCoV-19/Switzerland/BS-ETHZ-330428/2020\|EPI_ISL_61 | ···················· | ···················· | ···················· | ···················· | ···················· | ···················· |
| P | 7682 | hCoV-19/Switzerland/BS-ETHZ-330429/2020\|EPI_ISL_61 | ···················· | ·A·················· | ···················· | ···················· | ···················· | ···················· |
| P | 7683 | hCoV-19/Switzerland/BS-ETHZ-330430/2020\|EPI_ISL_61 | ···················· | ···················· | ···················· | ···················· | ···················· | ···················· |
| P | 7684 | hCoV-19/Switzerland/BS-ETHZ-330431/2020\|EPI_ISL_61 | ···················· | ···················· | ···················· | ···················· | ···················· | ···················· |
| P | 7685 | hCoV-19/Switzerland/BS-ETHZ-330433/2020\|EPI_ISL_61 | ···················· | ···················· | ···················· | ···················· | ···················· | ···················· |
| P | 7686 | hCoV-19/Switzerland/BE-ETHZ-330250/2020\|EPI_ISL_61 | ···················· | ···················· | ···················· | ···················· | ···················· | ···················· |
| P | 7687 | hCoV-19/Switzerland/BE-ETHZ-330421/2020\|EPI_ISL_61 | ···················· | ···················· | ···················· | ···················· | ···················· | ···················· |

| | | | | | | | |
|---|---|---|---|---|---|---|---|
| P | 7688 hCoV-19/Switzerland/BE-ETHZ-330423/2020\|EPI_ISL_61 | ···················· | ···················· | ···················· | ···················· | ···················· | ···················· |
| P | 7689 hCoV-19/Switzerland/BE-ETHZ-330513/2020\|EPI_ISL_61 | ···················· | ···················· | ···················· | ···················· | ···················· | ···················· |
| P | 7690 hCoV-19/Switzerland/BE-ETHZ-330514/2020\|EPI_ISL_61 | ···················· | ···················· | ···················· | ···················· | ···················· | ···················· |
| P | 7691 hCoV-19/Switzerland/BE-ETHZ-330515/2020\|EPI_ISL_61 | ···················· | ···················· | ···················· | ···················· | ···················· | ···················· |
| P | 7692 hCoV-19/Switzerland/BE-ETHZ-330516/2020\|EPI_ISL_61 | ···················· | ···················· | ···················· | ···················· | ···················· | ···················· |
| P | 7693 hCoV-19/Switzerland/BE-ETHZ-330517/2020\|EPI_ISL_61 | ···················· | ···················· | ···················· | ···················· | ···················· | ···················· |
| P | 7694 hCoV-19/Switzerland/BE-ETHZ-330518/2020\|EPI_ISL_61 | ···················· | ···················· | ···················· | ···················· | ···················· | ···················· |
| P | 7695 hCoV-19/Switzerland/BE-ETHZ-331067/2020\|EPI_ISL_61 | ···················· | ···················· | ···················· | ···················· | ···················· | ···················· |
| P | 7696 hCoV-19/Switzerland/BE-ETHZ-331070/2020\|EPI_ISL_61 | ···················· | ···················· | ···················· | ···················· | ···················· | ···················· |
| P | 7697 hCoV-19/Switzerland/BE-ETHZ-331078/2020\|EPI_ISL_61 | ···················· | ···················· | ···················· | ···················· | ···················· | ···················· |
| P | 7698 hCoV-19/Switzerland/BE-ETHZ-331081/2020\|EPI_ISL_61 | ···················· | ···················· | ···················· | ···················· | ···················· | ···················· |
| P | 7699 hCoV-19/Switzerland/BE-ETHZ-331447/2020\|EPI_ISL_61 | ···················· | ···················· | ···················· | ···················· | ···················· | ···················· |
| P | 7700 hCoV-19/Switzerland/BE-ETHZ-331456/2020\|EPI_ISL_61 | ···················· | ···················· | ···················· | ···················· | ···················· | ···················· |
| P | 7701 hCoV-19/Switzerland/BE-ETHZ-332173/2020\|EPI_ISL_61 | ···················· | ···················· | ···················· | ···················· | ···················· | ···················· |
| P | 7702 hCoV-19/Switzerland/BE-ETHZ-332176/2020\|EPI_ISL_61 | ···················· | ····G··············· | ···················· | ···················· | ···················· | ···················· |
| P | 7703 hCoV-19/Switzerland/BE-ETHZ-332178/2020\|EPI_ISL_61 | ···················· | ···················· | ···················· | ···················· | ···················· | ···················· |
| P | 7704 hCoV-19/Switzerland/BE-ETHZ-332199/2020\|EPI_ISL_61 | ···················· | ···················· | ···················· | ···················· | ···················· | ···················· |
| P | 7705 hCoV-19/Switzerland/GE-ETHZ-330055/2020\|EPI_ISL_61 | ···················· | ···················· | ···················· | ···················· | ···················· | ···················· |
| P | 7706 hCoV-19/Switzerland/VD-ETHZ-330253/2020\|EPI_ISL_61 | ···················· | ···················· | ···················· | ···················· | ···················· | ···················· |
| P | 7707 hCoV-19/Switzerland/VD-ETHZ-330254/2020\|EPI_ISL_61 | ···················· | ···················· | ···················· | ···················· | ···················· | ···················· |
| P | 7708 hCoV-19/Switzerland/VD-ETHZ-330255/2020\|EPI_ISL_61 | ···················· | ···················· | ···················· | ···················· | ···················· | ···················· |
| P | 7709 hCoV-19/Switzerland/VD-ETHZ-330257/2020\|EPI_ISL_61 | ···················· | ···················· | ···················· | ···················· | ···················· | ···················· |
| P | 7710 hCoV-19/Switzerland/VD-ETHZ-330258/2020\|EPI_ISL_61 | ···················· | ···················· | ···················· | ···················· | ···················· | ···················· |
| P | 7711 hCoV-19/Switzerland/VD-ETHZ-330259/2020\|EPI_ISL_61 | ···················· | ···················· | ···················· | ···················· | ···················· | ···················· |
| P | 7712 hCoV-19/Switzerland/VD-ETHZ-332170/2020\|EPI_ISL_61 | ···················· | ···················· | ···················· | ···················· | ···················· | ···················· |
| P | 7713 hCoV-19/Switzerland/ZH-ETHZ-330256/2020\|EPI_ISL_61 | ···················· | ···················· | ···················· | ···················· | ···················· | ···················· |
| P | 7714 hCoV-19/Switzerland/ZH-ETHZ-331069/2020\|EPI_ISL_61 | ···················· | ···················· | ···················· | ···················· | ···················· | ···················· |
| P | 7715 hCoV-19/Switzerland/ZH-ETHZ-331071/2020\|EPI_ISL_61 | ···················· | ···················· | ···················· | ···················· | ···················· | ···················· |
| P | 7716 hCoV-19/Switzerland/ZH-ETHZ-331072/2020\|EPI_ISL_61 | ···················· | ···················· | ···················· | ···················· | ···················· | ···················· |
| P | 7717 hCoV-19/Switzerland/ZH-ETHZ-331073/2020\|EPI_ISL_61 | ···················· | ···················· | ···················· | ···················· | ···················· | ···················· |
| P | 7718 hCoV-19/Switzerland/ZH-ETHZ-331074/2020\|EPI_ISL_61 | ···················· | ···················· | ···················· | ···················· | ···················· | ···················· |
| P | 7719 hCoV-19/Switzerland/ZH-ETHZ-331075/2020\|EPI_ISL_61 | ···················· | ···················· | ···················· | ···················· | ···················· | ···················· |
| P | 7720 hCoV-19/Switzerland/ZH-ETHZ-331077/2020\|EPI_ISL_61 | ···················· | ···················· | ···················· | ···················· | ···················· | ···················· |
| P | 7721 hCoV-19/Switzerland/ZH-ETHZ-331079/2020\|EPI_ISL_61 | ···················· | ···················· | ···················· | ···················· | ···················· | ···················· |
| P | 7722 hCoV-19/Switzerland/ZH-ETHZ-331451/2020\|EPI_ISL_61 | ···················· | ···················· | ···················· | ···················· | ···················· | ···················· |
| P | 7723 hCoV-19/Switzerland/ZH-ETHZ-331459/2020\|EPI_ISL_61 | ···················· | ···················· | ···················· | ···················· | ······K············· | ···················· |
| P | 7724 hCoV-19/Switzerland/ZH-ETHZ-331460/2020\|EPI_ISL_61 | ···················· | ···················· | ···················· | ···················· | ···················· | ···················· |
| P | 7725 hCoV-19/Switzerland/ZH-ETHZ-331463/2020\|EPI_ISL_61 | ···················· | ···················· | ···················· | ···················· | ···················· | ···················· |
| P | 7726 hCoV-19/Switzerland/ZH-ETHZ-332169/2020\|EPI_ISL_61 | ···················· | ···················· | ···················· | ···················· | ···················· | ···················· |
| P | 7727 hCoV-19/Switzerland/ZH-ETHZ-332177/2020\|EPI_ISL_61 | ···················· | ····A··············· | ···················· | ···················· | ···················· | ···················· |
| P | 7728 hCoV-19/Gibraltar/204462807/2020\|EPI_ISL_614982\|20 | ···················· | ···················· | ···················· | ···················· | ···················· | ···················· |
| P | 7729 hCoV-19/Gibraltar/204462808/2020\|EPI_ISL_614983\|20 | ···················· | ···················· | ···················· | ···················· | ···················· | ···················· |
| P | 7730 hCoV-19/Gibraltar/204462809/2020\|EPI_ISL_614984\|20 | ···················· | ···················· | ···················· | ···················· | ···················· | ···················· |
| P | 7731 hCoV-19/Gibraltar/204462810/2020\|EPI_ISL_614985\|20 | ···················· | ···················· | ···················· | ···················· | ···················· | ···················· |
| P | 7732 hCoV-19/Gibraltar/204462811/2020\|EPI_ISL_614986\|20 | ···················· | ···················· | ···················· | ···················· | ···················· | ···················· |
| P | 7733 hCoV-19/Gibraltar/204462812/2020\|EPI_ISL_614987\|20 | ···················· | ···················· | ···················· | ···················· | ···················· | ···················· |
| P | 7734 hCoV-19/Gibraltar/204462813/2020\|EPI_ISL_614988\|20 | ···················· | ···················· | ···················· | ···················· | ···················· | ···················· |
| P | 7735 hCoV-19/Gibraltar/204462814/2020\|EPI_ISL_614989\|20 | ···················· | ···················· | ···················· | ···················· | ···················· | ···················· |
| P | 7736 hCoV-19/Gibraltar/204462815/2020\|EPI_ISL_614990\|20 | ···················· | ···················· | ···················· | ···················· | ···················· | ···················· |
| P | 7737 hCoV-19/Gibraltar/204462816/2020\|EPI_ISL_614991\|20 | ···················· | ···················· | ···················· | ···················· | ···················· | ···················· |
| P | 7738 hCoV-19/Gibraltar/204462817/2020\|EPI_ISL_614992\|20 | ···················· | ···················· | ···················· | ···················· | ···················· | ···················· |
| P | 7739 hCoV-19/Gibraltar/204462818/2020\|EPI_ISL_614993\|20 | ···················· | ···················· | ···················· | ···················· | ···················· | ···················· |
| P | 7740 hCoV-19/Gibraltar/204462819/2020\|EPI_ISL_614994\|20 | ···················· | ···················· | ···················· | ···················· | ···················· | ···················· |
| P | 7741 hCoV-19/Gibraltar/204462822/2020\|EPI_ISL_614997\|20 | ···················· | ···················· | ···················· | ···················· | ···················· | ···················· |
| P | 7742 hCoV-19/Gibraltar/204462823/2020\|EPI_ISL_614998\|20 | ···················· | ···················· | ···················· | ···················· | ···················· | ···················· |
| P | 7743 hCoV-19/Gibraltar/204462824/2020\|EPI_ISL_614999\|20 | ···················· | ···················· | ···················· | ···················· | ···················· | ···················· |
| P | 7744 hCoV-19/Denmark/DCGC-7989/2020\|EPI_ISL_617110\|2020 | ···················· | ·A·················· | ···················· | ···················· | ···················· | ···················· |
| P | 7745 hCoV-19/Denmark/DCGC-8000/2020\|EPI_ISL_617111\|2020 | ···················· | ···················· | ···················· | ···················· | ···················· | ···················· |
| P | 7746 hCoV-19/Denmark/DCGC-8005/2020\|EPI_ISL_617112\|2020 | ···················· | ···················· | ···················· | ···················· | ···················· | ···················· |
| P | 7747 hCoV-19/Denmark/DCGC-7971/2020\|EPI_ISL_617113\|2020 | ···················· | ···················· | ···················· | ···················· | ···················· | ···················· |
| P | 7748 hCoV-19/Denmark/DCGC-7977/2020\|EPI_ISL_617114\|2020 | ···················· | ···················· | ···················· | ···················· | ···················· | ···················· |
| P | 7749 hCoV-19/Denmark/DCGC-8001/2020\|EPI_ISL_617115\|2020 | ···················· | ····A··············· | ···················· | ···················· | ···················· | ···················· |
| P | 7750 hCoV-19/Denmark/DCGC-8006/2020\|EPI_ISL_617116\|2020 | ···················· | ···················· | ···················· | ···················· | ···················· | ···················· |
| P | 7751 hCoV-19/Denmark/DCGC-8013/2020\|EPI_ISL_617117\|2020 | ···················· | ···················· | ···················· | ···················· | ···················· | ···················· |
| P | 7752 hCoV-19/Denmark/DCGC-8031/2020\|EPI_ISL_617118\|2020 | ···················· | ···················· | ···················· | ···················· | ···················· | ···················· |
| P | 7753 hCoV-19/Denmark/DCGC-7972/2020\|EPI_ISL_617119\|2020 | ···················· | ···················· | ···················· | ···················· | ···················· | ···················· |
| P | 7754 hCoV-19/Denmark/DCGC-7978/2020\|EPI_ISL_617120\|2020 | ···················· | ···················· | ···················· | ···················· | ···················· | ···················· |
| P | 7755 hCoV-19/Denmark/DCGC-7984/2020\|EPI_ISL_617121\|2020 | ···················· | ···················· | ···················· | ···················· | ···················· | ···················· |
| P | 7756 hCoV-19/Denmark/DCGC-8007/2020\|EPI_ISL_617122\|2020 | ···················· | ···················· | ···················· | ···················· | ···················· | ···················· |
| P | 7757 hCoV-19/Denmark/DCGC-8014/2020\|EPI_ISL_617123\|2020 | ···················· | ···················· | ···················· | ···················· | ···················· | ···················· |
| P | 7758 hCoV-19/Denmark/DCGC-8032/2020\|EPI_ISL_617124\|2020 | ···················· | ···················· | ···················· | ···················· | ···················· | ···················· |
| P | 7759 hCoV-19/Denmark/DCGC-8039/2020\|EPI_ISL_617125\|2020 | ···················· | ···················· | ···················· | ···················· | ···················· | ···················· |
| P | 7760 hCoV-19/Denmark/DCGC-7973/2020\|EPI_ISL_617126\|2020 | ···················· | ···················· | ···················· | ···················· | ···················· | ···················· |
| P | 7761 hCoV-19/Denmark/DCGC-7990/2020\|EPI_ISL_617127\|2020 | ···················· | ···················· | ···················· | ···················· | ···················· | ···················· |
| P | 7762 hCoV-19/Denmark/DCGC-7995/2020\|EPI_ISL_617128\|2020 | ···················· | ···················· | ···················· | ···················· | ···················· | ···················· |
| P | 7763 hCoV-19/Denmark/DCGC-8008/2020\|EPI_ISL_617129\|2020 | ···················· | ···················· | ···················· | ···················· | ···················· | ···················· |
| P | 7764 hCoV-19/Denmark/DCGC-8015/2020\|EPI_ISL_617130\|2020 | ···················· | ·A·················· | ···················· | ···················· | ···················· | ···················· |
| P | 7765 hCoV-19/Denmark/DCGC-8019/2020\|EPI_ISL_617131\|2020 | ···················· | ···················· | ···················· | ···················· | ···················· | ···················· |
| P | 7766 hCoV-19/Denmark/DCGC-8033/2020\|EPI_ISL_617132\|2020 | ···················· | ···················· | ···················· | ···················· | ···················· | ···················· |
| P | 7767 hCoV-19/Denmark/DCGC-7991/2020\|EPI_ISL_617133\|2020 | ···················· | ···················· | ···················· | ···················· | ···················· | ···················· |

| | | | | | | | |
|---|---|---|---|---|---|---|---|
| P | 7768 hCoV-19/Denmark/DCGC-7996/2020\|EPI_ISL_617134\|2020 | ···················· | ···················· | ····················· | ···················· | ···················· | ···················· |
| P | 7769 hCoV-19/Denmark/DCGC-8009/2020\|EPI_ISL_617135\|2020 | ···················· | ···················· | ····················· | ···················· | ···················· | ···················· |
| P | 7770 hCoV-19/Denmark/DCGC-8016/2020\|EPI_ISL_617136\|2020 | ···················· | ···················· | ····················· | ···················· | ···················· | ···················· |
| P | 7771 hCoV-19/Denmark/DCGC-7980/2020\|EPI_ISL_617137\|2020 | ···················· | ···················· | ····················· | ···················· | ···················· | ···················· |
| P | 7772 hCoV-19/Denmark/DCGC-7997/2020\|EPI_ISL_617138\|2020 | ···················· | ·A·················· | ····················· | ···················· | ···················· | ···················· |
| P | 7773 hCoV-19/Denmark/DCGC-8027/2020\|EPI_ISL_617139\|2020 | ···················· | ···················· | ····················· | ···················· | ···················· | ···················· |
| P | 7774 hCoV-19/Denmark/DCGC-8034/2020\|EPI_ISL_617140\|2020 | ···················· | ···················· | ····················· | ···················· | ···················· | ···················· |
| P | 7775 hCoV-19/Denmark/DCGC-7981/2020\|EPI_ISL_617141\|2020 | ···················· | ···················· | ····················· | ···················· | ···················· | ···················· |
| P | 7776 hCoV-19/Denmark/DCGC-7988/2020\|EPI_ISL_617142\|2020 | ···················· | ···················· | ····················· | ···················· | ···················· | ···················· |
| P | 7777 hCoV-19/Denmark/DCGC-8004/2020\|EPI_ISL_617143\|2020 | ···················· | ···················· | ····················· | ···················· | ···················· | ···················· |
| P | 7778 hCoV-19/Denmark/DCGC-8021/2020\|EPI_ISL_617144\|2020 | ···················· | ···················· | ····················· | ···················· | ···················· | ···················· |
| P | 7779 hCoV-19/Denmark/DCGC-8035/2020\|EPI_ISL_617145\|2020 | ···················· | ···················· | ····················· | ···················· | ···················· | ···················· |
| P | 7780 hCoV-19/Denmark/DCGC-7975/2020\|EPI_ISL_617146\|2020 | ···················· | ···················· | ····················· | ···················· | ···················· | ···················· |
| P | 7781 hCoV-19/Denmark/DCGC-7982/2020\|EPI_ISL_617147\|2020 | ···················· | ···················· | ····················· | ···················· | ···················· | ···················· |
| P | 7782 hCoV-19/Denmark/DCGC-7993/2020\|EPI_ISL_617148\|2020 | ···················· | ···················· | ····················· | ···················· | ···················· | ···················· |
| P | 7783 hCoV-19/Denmark/DCGC-8012/2020\|EPI_ISL_617149\|2020 | ···················· | ···················· | ····················· | ···················· | ···················· | ···················· |
| P | 7784 hCoV-19/Denmark/DCGC-8054/2020\|EPI_ISL_617180\|2020 | ···················· | ···················· | ····················· | ···················· | ···················· | ···················· |
| P | 7785 hCoV-19/Denmark/DCGC-8061/2020\|EPI_ISL_617181\|2020 | ···················· | ···················· | ····················· | ···················· | ···················· | ···················· |
| P | 7786 hCoV-19/Denmark/DCGC-8091/2020\|EPI_ISL_617182\|2020 | ···················· | ···················· | ····················· | ···················· | ···················· | ···················· |
| P | 7787 hCoV-19/Denmark/DCGC-8100/2020\|EPI_ISL_617183\|2020 | ···················· | ···················· | ····················· | ···················· | ···················· | ···················· |
| P | 7788 hCoV-19/Denmark/DCGC-8105/2020\|EPI_ISL_617184\|2020 | ···················· | ···················· | ····················· | ···················· | ···················· | ···················· |
| P | 7789 hCoV-19/Denmark/DCGC-8081/2020\|EPI_ISL_617188\|2020 | ···················· | ···················· | ····················· | ···················· | ···················· | ···················· |
| P | 7790 hCoV-19/Denmark/DCGC-8092/2020\|EPI_ISL_617190\|2020 | ···················· | ···················· | ····················· | ···················· | ···················· | ···················· |
| P | 7791 hCoV-19/Denmark/DCGC-8094/2020\|EPI_ISL_617191\|2020 | ···················· | ···················· | ····················· | ···················· | ···················· | ···················· |
| P | 7792 hCoV-19/Denmark/DCGC-8106/2020\|EPI_ISL_617192\|2020 | ···················· | ···················· | ····················· | ···················· | ···················· | ···················· |
| P | 7793 hCoV-19/Denmark/DCGC-8063/2020\|EPI_ISL_617193\|2020 | ···················· | ···················· | ····················· | ···················· | ···················· | ···················· |
| P | 7794 hCoV-19/Denmark/DCGC-8076/2020\|EPI_ISL_617194\|2020 | ···················· | ···················· | ····················· | ···················· | ···················· | ···················· |
| P | 7795 hCoV-19/Denmark/DCGC-8082/2020\|EPI_ISL_617195\|2020 | ···················· | ···················· | ····················· | ···················· | ···················· | ···················· |
| P | 7796 hCoV-19/Denmark/DCGC-8087/2020\|EPI_ISL_617196\|2020 | ···················· | ···················· | ····················· | ···················· | ···················· | ···················· |
| P | 7797 hCoV-19/Denmark/DCGC-8095/2020\|EPI_ISL_617197\|2020 | ···················· | ···················· | ····················· | ···················· | ···················· | ···················· |
| P | 7798 hCoV-19/Denmark/DCGC-8101/2020\|EPI_ISL_617198\|2020 | ···················· | ·A·················· | ····················· | ···················· | ···················· | ···················· |
| P | 7799 hCoV-19/Denmark/DCGC-8125/2020\|EPI_ISL_617199\|2020 | ···················· | ·A·················· | ····················· | ···················· | ···················· | ···A················ |
| P | 7800 hCoV-19/Denmark/DCGC-7578/2020\|EPI_ISL_618440\|2020 | ···················· | ···················· | ····················· | ···················· | ···················· | ···················· |
| P | 7801 hCoV-19/Denmark/DCGC-7528/2020\|EPI_ISL_618442\|2020 | ···················· | ·A·················· | ····················· | ···················· | ···················· | ···A················ |
| P | 7802 hCoV-19/Denmark/DCGC-7536/2020\|EPI_ISL_618443\|2020 | ···················· | ···················· | ····················· | ···················· | ···················· | ···················· |
| P | 7803 hCoV-19/Denmark/DCGC-7540/2020\|EPI_ISL_618444\|2020 | ···················· | ···················· | ····················· | ···················· | ···················· | ···················· |
| P | 7804 hCoV-19/Denmark/DCGC-7559/2020\|EPI_ISL_618446\|2020 | ···················· | ···················· | ····················· | ···················· | ···················· | ···················· |
| P | 7805 hCoV-19/Denmark/DCGC-7567/2020\|EPI_ISL_618447\|2020 | ···················· | ···················· | ····················· | ···················· | ···················· | ···················· |
| P | 7806 hCoV-19/Denmark/DCGC-7572/2020\|EPI_ISL_618448\|2020 | ···················· | ···················· | ····················· | ···················· | ···················· | ···················· |
| P | 7807 hCoV-19/Denmark/DCGC-7584/2020\|EPI_ISL_618449\|2020 | ···················· | ···················· | ····················· | ···················· | ···················· | ···················· |
| P | 7808 hCoV-19/Denmark/DCGC-7596/2020\|EPI_ISL_618450\|2020 | ···················· | ···················· | ····················· | ···················· | ···················· | ···················· |
| P | 7809 hCoV-19/Denmark/DCGC-7529/2020\|EPI_ISL_618451\|2020 | ···················· | ···················· | ····················· | ···················· | ···················· | ···················· |
| P | 7810 hCoV-19/Denmark/DCGC-7537/2020\|EPI_ISL_618452\|2020 | ···················· | ···················· | ····················· | ···················· | ···················· | ···················· |
| P | 7811 hCoV-19/Denmark/DCGC-7541/2020\|EPI_ISL_618453\|2020 | ···················· | ···················· | ····················· | ···················· | ···················· | ···················· |
| P | 7812 hCoV-19/Denmark/DCGC-7548/2020\|EPI_ISL_618454\|2020 | ···················· | ···················· | ····················· | ···················· | ···················· | ···················· |
| P | 7813 hCoV-19/Denmark/DCGC-7555/2020\|EPI_ISL_618455\|2020 | ···················· | ···················· | ····················· | ···················· | ···················· | ···················· |
| P | 7814 hCoV-19/Denmark/DCGC-7560/2020\|EPI_ISL_618456\|2020 | ···················· | ···················· | ····················· | ···················· | ···················· | ···················· |
| P | 7815 hCoV-19/Denmark/DCGC-7573/2020\|EPI_ISL_618457\|2020 | ···················· | ···················· | ····················· | ···················· | ···················· | ···················· |
| P | 7816 hCoV-19/Denmark/DCGC-7579/2020\|EPI_ISL_618458\|2020 | ···················· | ···················· | ····················· | ···················· | ···················· | ···················· |
| P | 7817 hCoV-19/Denmark/DCGC-7585/2020\|EPI_ISL_618459\|2020 | ···················· | ···················· | ····················· | ···················· | ···················· | ···················· |
| P | 7818 hCoV-19/Denmark/DCGC-7591/2020\|EPI_ISL_618460\|2020 | ···················· | ···················· | ····················· | ···················· | ···················· | ···················· |
| P | 7819 hCoV-19/Denmark/DCGC-7597/2020\|EPI_ISL_618461\|2020 | ···················· | ···················· | ····················· | ···················· | ···················· | ···················· |
| P | 7820 hCoV-19/Denmark/DCGC-7530/2020\|EPI_ISL_618462\|2020 | ···················· | ···················· | ····················· | ···················· | ···················· | ···················· |
| P | 7821 hCoV-19/Denmark/DCGC-7542/2020\|EPI_ISL_618463\|2020 | ···················· | ···················· | ····················· | ···················· | ···················· | ···················· |
| P | 7822 hCoV-19/Denmark/DCGC-7556/2020\|EPI_ISL_618465\|2020 | ···················· | ···················· | ····················· | ···················· | ···················· | ···················· |
| P | 7823 hCoV-19/Denmark/DCGC-7568/2020\|EPI_ISL_618467\|2020 | ···················· | ···················· | ····················· | ···················· | ···················· | ···················· |
| P | 7824 hCoV-19/Denmark/DCGC-7580/2020\|EPI_ISL_618468\|2020 | ···················· | ···················· | ····················· | ···················· | ···················· | ···················· |
| P | 7825 hCoV-19/Denmark/DCGC-7592/2020\|EPI_ISL_618470\|2020 | ···················· | ···················· | ····················· | ···················· | ···················· | ···················· |
| P | 7826 hCoV-19/Denmark/DCGC-7598/2020\|EPI_ISL_618471\|2020 | ···················· | ·A·················· | ····················· | ···················· | ···················· | ···················· |
| P | 7827 hCoV-19/Denmark/DCGC-7557/2020\|EPI_ISL_618475\|2020 | ···················· | ···················· | ····················· | ···················· | ···················· | ···················· |
| P | 7828 hCoV-19/Denmark/DCGC-7562/2020\|EPI_ISL_618476\|2020 | ···················· | ···················· | ····················· | ···················· | ······T············· | ···················· |
| P | 7829 hCoV-19/Denmark/DCGC-7574/2020\|EPI_ISL_618477\|2020 | ···················· | ···················· | ····················· | ···················· | ···················· | ···················· |
| P | 7830 hCoV-19/Denmark/DCGC-7587/2020\|EPI_ISL_618478\|2020 | ···················· | ···················· | ····················· | ···················· | ···················· | ···················· |
| P | 7831 hCoV-19/Denmark/DCGC-1677/2020\|EPI_ISL_618479\|2020 | ···················· | ···················· | ····················· | ···················· | ···················· | ···················· |
| P | 7832 hCoV-19/Denmark/DCGC-6925/2020\|EPI_ISL_619771\|2020 | ···················· | ···················· | ····················· | ···················· | ···················· | ···················· |
| P | 7833 hCoV-19/Denmark/DCGC-6926/2020\|EPI_ISL_619772\|2020 | ···················· | ···················· | ····················· | ···················· | ···················· | ···················· |
| P | 7834 hCoV-19/Denmark/DCGC-6927/2020\|EPI_ISL_619773\|2020 | ···················· | ·A·················· | ····················· | ···················· | ···················· | ···A················ |
| P | 7835 hCoV-19/Denmark/DCGC-6928/2020\|EPI_ISL_619774\|2020 | ···················· | ·A·················· | ····················· | ···················· | ···················· | ···A················ |
| P | 7836 hCoV-19/Denmark/DCGC-6929/2020\|EPI_ISL_619775\|2020 | ···················· | ·A·················· | ····················· | ···················· | ···················· | ···A················ |
| P | 7837 hCoV-19/Denmark/DCGC-6930/2020\|EPI_ISL_619776\|2020 | ···················· | ·A·················· | ····················· | ···················· | ···················· | ···A················ |
| P | 7838 hCoV-19/Denmark/DCGC-6932/2020\|EPI_ISL_619778\|2020 | ···················· | ···················· | ····················· | ···················· | ···················· | ···················· |
| P | 7839 hCoV-19/Denmark/DCGC-6933/2020\|EPI_ISL_619779\|2020 | ···················· | ···················· | ····················· | ···················· | ···················· | ···················· |
| P | 7840 hCoV-19/Denmark/DCGC-6934/2020\|EPI_ISL_619780\|2020 | ···················· | ···················· | ····················· | ···················· | ···················· | ···················· |
| P | 7841 hCoV-19/Denmark/DCGC-6935/2020\|EPI_ISL_619781\|2020 | ···················· | ·A·················· | ····················· | ···················· | ···················· | ···A················ |
| P | 7842 hCoV-19/Denmark/DCGC-6936/2020\|EPI_ISL_619782\|2020 | ···················· | ···················· | ····················· | ···················· | ···················· | ···················· |
| P | 7843 hCoV-19/Denmark/DCGC-6937/2020\|EPI_ISL_619783\|2020 | ···················· | ···················· | ····················· | ···················· | ···················· | ···················· |
| P | 7844 hCoV-19/Denmark/DCGC-6938/2020\|EPI_ISL_619784\|2020 | ···················· | ···················· | ····················· | ···················· | ···················· | ···················· |
| P | 7845 hCoV-19/Denmark/DCGC-6939/2020\|EPI_ISL_619785\|2020 | ···················· | ···················· | ····················· | ···················· | ···················· | ···················· |
| P | 7846 hCoV-19/Denmark/DCGC-6940/2020\|EPI_ISL_619786\|2020 | ···················· | ···················· | ····················· | ···················· | ···················· | ···················· |
| P | 7847 hCoV-19/Denmark/DCGC-6941/2020\|EPI_ISL_619787\|2020 | ···················· | ···················· | ····················· | ···················· | ···················· | ···················· |

| | | | | | | | | |
|---|---|---|---|---|---|---|---|---|
| P | 7848 | hCoV-19/Denmark/DCGC-6943/2020\|EPI_ISL_619789\|2020 | ···················· | ···················· | ···················· | ···················· | ···················· | ···················· |
| P | 7849 | hCoV-19/Denmark/DCGC-6945/2020\|EPI_ISL_619791\|2020 | ···················· | ···················· | ···················· | ···················· | ···················· | ···················· |
| P | 7850 | hCoV-19/Denmark/DCGC-6946/2020\|EPI_ISL_619792\|2020 | ···················· | ·A·················· | ···················· | ···················· | ···················· | ··A················· |
| P | 7851 | hCoV-19/Denmark/DCGC-6947/2020\|EPI_ISL_619793\|2020 | ···················· | ···················· | ···················· | ···················· | ···················· | ···················· |
| P | 7852 | hCoV-19/Denmark/DCGC-6948/2020\|EPI_ISL_619794\|2020 | ···················· | ···················· | ···················· | ···················· | ···················· | ···················· |
| P | 7853 | hCoV-19/Denmark/DCGC-6949/2020\|EPI_ISL_619795\|2020 | ···················· | ···················· | ···················· | ···················· | ···················· | ···················· |
| P | 7854 | hCoV-19/Denmark/DCGC-6950/2020\|EPI_ISL_619796\|2020 | ···················· | ···················· | ···················· | ···················· | ···················· | ···················· |
| P | 7855 | hCoV-19/Denmark/DCGC-6952/2020\|EPI_ISL_619798\|2020 | ···················· | ···················· | ···················· | ···················· | ···················· | ···················· |
| P | 7856 | hCoV-19/Denmark/DCGC-6953/2020\|EPI_ISL_619799\|2020 | ···················· | ···················· | ···················· | ···················· | ···················· | ···················· |
| P | 7857 | hCoV-19/Denmark/DCGC-5224/2020\|EPI_ISL_620762\|2020 | ···················· | ···················· | ···················· | ···················· | ···················· | ···················· |
| P | 7858 | hCoV-19/Denmark/DCGC-5438/2020\|EPI_ISL_620763\|2020 | ···················· | ···················· | ···················· | ···················· | ···················· | ···················· |
| P | 7859 | hCoV-19/Denmark/DCGC-5439/2020\|EPI_ISL_620764\|2020 | ···················· | ···················· | ···················· | ···················· | ···················· | ···················· |
| P | 7860 | hCoV-19/Denmark/DCGC-5440/2020\|EPI_ISL_620765\|2020 | ···················· | ···················· | ···················· | ···················· | ···················· | ···················· |
| P | 7861 | hCoV-19/Denmark/DCGC-5441/2020\|EPI_ISL_620766\|2020 | ···················· | ···················· | ···················· | ···················· | ···················· | ···················· |
| P | 7862 | hCoV-19/Denmark/DCGC-5442/2020\|EPI_ISL_620767\|2020 | ···················· | ···················· | ···················· | ···················· | ···················· | ···················· |
| P | 7863 | hCoV-19/Denmark/DCGC-5443/2020\|EPI_ISL_620768\|2020 | ···················· | ···················· | ···················· | ···················· | ···················· | ···················· |
| P | 7864 | hCoV-19/Denmark/DCGC-5444/2020\|EPI_ISL_620769\|2020 | ···················· | ···················· | ···················· | ···················· | ···················· | ···················· |
| P | 7865 | hCoV-19/Denmark/DCGC-5445/2020\|EPI_ISL_620770\|2020 | ···················· | ···················· | ···················· | ···················· | ···················· | ···················· |
| P | 7866 | hCoV-19/Denmark/DCGC-5446/2020\|EPI_ISL_620771\|2020 | ···················· | ···················· | ···················· | ···················· | ···················· | ···················· |
| P | 7867 | hCoV-19/Denmark/DCGC-5447/2020\|EPI_ISL_620772\|2020 | ···················· | ···················· | ···················· | ···················· | ···················· | ···················· |
| P | 7868 | hCoV-19/Denmark/DCGC-5448/2020\|EPI_ISL_620773\|2020 | ···················· | ···················· | ···················· | ···················· | ···················· | ···················· |
| P | 7869 | hCoV-19/Denmark/DCGC-5449/2020\|EPI_ISL_620774\|2020 | ···················· | ···················· | ···················· | ···················· | ···················· | ···················· |
| P | 7870 | hCoV-19/Denmark/DCGC-5450/2020\|EPI_ISL_620775\|2020 | ···················· | ···················· | ···················· | ···················· | ···················· | ···················· |
| P | 7871 | hCoV-19/Denmark/DCGC-5451/2020\|EPI_ISL_620776\|2020 | ···················· | ···················· | ···················· | ···················· | ···················· | ···················· |
| P | 7872 | hCoV-19/Denmark/DCGC-5452/2020\|EPI_ISL_620777\|2020 | ···················· | ···················· | ···················· | ···················· | ···················· | ···················· |
| P | 7873 | hCoV-19/Denmark/DCGC-5453/2020\|EPI_ISL_620778\|2020 | ···················· | ···················· | ···················· | ···················· | ···················· | ···················· |
| P | 7874 | hCoV-19/Denmark/DCGC-5455/2020\|EPI_ISL_620780\|2020 | ···················· | ···················· | ···················· | ···················· | ···················· | ···················· |
| P | 7875 | hCoV-19/Denmark/DCGC-5456/2020\|EPI_ISL_620781\|2020 | ···················· | ···················· | ···················· | ···················· | ···················· | ···················· |
| P | 7876 | hCoV-19/Denmark/DCGC-5457/2020\|EPI_ISL_620782\|2020 | ···················· | ···················· | ···················· | ···················· | ···················· | ···················· |
| P | 7877 | hCoV-19/Denmark/DCGC-5458/2020\|EPI_ISL_620783\|2020 | ···················· | ···················· | ···················· | ···················· | ···················· | ···················· |
| P | 7878 | hCoV-19/Denmark/DCGC-5459/2020\|EPI_ISL_620784\|2020 | ···················· | ···················· | ···················· | ···················· | ···················· | ···················· |
| P | 7879 | hCoV-19/Denmark/DCGC-5460/2020\|EPI_ISL_620785\|2020 | ···················· | ···················· | ···················· | ···················· | ···················· | ···················· |
| P | 7880 | hCoV-19/Denmark/DCGC-5461/2020\|EPI_ISL_620786\|2020 | ···················· | ···················· | ···················· | ···················· | ···················· | ···················· |
| P | 7881 | hCoV-19/Denmark/DCGC-5462/2020\|EPI_ISL_620787\|2020 | ···················· | ···················· | ···················· | ···················· | ···················· | ···················· |
| P | 7882 | hCoV-19/Denmark/DCGC-5463/2020\|EPI_ISL_620788\|2020 | ···················· | ···················· | ···················· | ···················· | ···················· | ···················· |
| P | 7883 | hCoV-19/Denmark/DCGC-5464/2020\|EPI_ISL_620789\|2020 | ···················· | ···················· | ···················· | ···················· | ·····T·············· | ···················· |
| P | 7884 | hCoV-19/Denmark/DCGC-5465/2020\|EPI_ISL_620790\|2020 | ···················· | ···················· | ···················· | ···················· | ···················· | ···················· |
| P | 7885 | hCoV-19/Denmark/DCGC-5466/2020\|EPI_ISL_620791\|2020 | ···················· | ···················· | ···················· | ···················· | ···················· | ···················· |
| P | 7886 | hCoV-19/Denmark/DCGC-5467/2020\|EPI_ISL_620792\|2020 | ···················· | ···················· | ···················· | ···················· | ···················· | ···················· |
| P | 7887 | hCoV-19/Denmark/DCGC-5468/2020\|EPI_ISL_620793\|2020 | ···················· | ···················· | ···················· | ···················· | ···················· | ···················· |
| P | 7888 | hCoV-19/Denmark/DCGC-5469/2020\|EPI_ISL_620794\|2020 | ···················· | ···················· | ···················· | ···················· | ···················· | ···················· |
| P | 7889 | hCoV-19/Denmark/DCGC-5470/2020\|EPI_ISL_620795\|2020 | ···················· | ···················· | ···················· | ···················· | ···················· | ···················· |
| P | 7890 | hCoV-19/Denmark/DCGC-5471/2020\|EPI_ISL_620796\|2020 | ···················· | ·A·················· | ···················· | ···················· | ···················· | ···················· |
| P | 7891 | hCoV-19/Denmark/DCGC-5472/2020\|EPI_ISL_620797\|2020 | ···················· | ···················· | ···················· | ···················· | ···················· | ···················· |
| P | 7892 | hCoV-19/England/ALDP-AA2060/2020\|EPI_ISL_624300\|20 | ···················· | ···················· | ···················· | ···················· | ···················· | ···················· |
| P | 7893 | hCoV-19/England/ALDP-AA4277/2020\|EPI_ISL_624301\|20 | ···················· | ···················· | ···················· | ···················· | ···················· | ···················· |
| P | 7894 | hCoV-19/England/ALDP-AA1C8B/2020\|EPI_ISL_624302\|20 | ···················· | ···················· | ···················· | ···················· | ···················· | ···················· |
| P | 7895 | hCoV-19/England/ALDP-AA1F91/2020\|EPI_ISL_624304\|20 | ···················· | ···················· | ···················· | ···················· | ···················· | ···················· |
| P | 7896 | hCoV-19/England/ALDP-AA281D/2020\|EPI_ISL_624305\|20 | ···················· | ···················· | ···················· | ···················· | ···················· | ···················· |
| P | 7897 | hCoV-19/England/ALDP-AA10AD/2020\|EPI_ISL_624306\|20 | ···················· | ···················· | ···················· | ···················· | ···················· | ···················· |
| P | 7898 | hCoV-19/England/ALDP-AA1D6A/2020\|EPI_ISL_624308\|20 | ···················· | ···················· | ···················· | ···················· | ···················· | ···················· |
| P | 7899 | hCoV-19/England/ALDP-AA3DB3/2020\|EPI_ISL_624309\|20 | ···················· | ···················· | ···················· | ···················· | ···················· | ···················· |
| P | 7900 | hCoV-19/England/ALDP-AA3C10/2020\|EPI_ISL_624310\|20 | ···················· | ···················· | ···················· | ···················· | ···················· | ···················· |
| P | 7901 | hCoV-19/England/ALDP-AA3BE6/2020\|EPI_ISL_624311\|20 | ···················· | ···················· | ···················· | ···················· | ···················· | ···················· |
| P | 7902 | hCoV-19/England/ALDP-AA1F19/2020\|EPI_ISL_624312\|20 | ···················· | ···················· | ···················· | ···················· | ···················· | ···················· |
| P | 7903 | hCoV-19/England/ALDP-AA3A52/2020\|EPI_ISL_624313\|20 | ···················· | ···················· | ···················· | ···················· | ···················· | ···················· |
| P | 7904 | hCoV-19/England/ALDP-AA1F28/2020\|EPI_ISL_624314\|20 | ···················· | ···················· | ···················· | ···················· | ·····T·············· | ···················· |
| P | 7905 | hCoV-19/England/ALDP-AA475A/2020\|EPI_ISL_624315\|20 | ···················· | ···················· | ···················· | ···················· | ···················· | ···················· |
| P | 7906 | hCoV-19/England/ALDP-AA3CC5/2020\|EPI_ISL_624316\|20 | ···················· | ···················· | ···················· | ···················· | ···················· | ···················· |
| P | 7907 | hCoV-19/England/ALDP-AA3E65/2020\|EPI_ISL_624317\|20 | ···················· | ···················· | ···················· | ···················· | ···················· | ···················· |
| P | 7908 | hCoV-19/USA/CA-CZB-11928/2020\|EPI_ISL_625605\|2020- | ···················· | ···················· | ···················· | ···················· | ···················· | ···················· |
| P | 7909 | hCoV-19/USA/CA-CZB-11929/2020\|EPI_ISL_625606\|2020- | ···················· | ···················· | ···················· | ···················· | ···················· | ···················· |
| P | 7910 | hCoV-19/USA/CA-CZB-11931/2020\|EPI_ISL_625607\|2020- | ···················· | ·A·················· | ···················· | ···················· | ···················· | ···················· |
| P | 7912 | hCoV-19/USA/CA-CZB-11937/2020\|EPI_ISL_625610\|2020- | ···················· | ·A·················· | ···················· | ···················· | ···················· | ···················· |
| P | 7913 | hCoV-19/USA/CA-CZB-11850/2020\|EPI_ISL_625611\|2020- | ···················· | ···················· | ···················· | ···················· | ···················· | ···················· |
| P | 7914 | hCoV-19/USA/CA-CZB-11852/2020\|EPI_ISL_625612\|2020- | ···················· | ·A·················· | ···················· | ···················· | ···················· | ···················· |
| P | 7915 | hCoV-19/USA/CA-CZB-11854/2020\|EPI_ISL_625613\|2020- | ···················· | ·A·················· | ···················· | ···················· | ···················· | ···················· |
| P | 7916 | hCoV-19/USA/CA-CZB-11855/2020\|EPI_ISL_625614\|2020- | ···················· | ···················· | ···················· | ···················· | ···················· | ···················· |
| P | 7917 | hCoV-19/USA/CA-CZB-11859/2020\|EPI_ISL_625616\|2020- | ···················· | ···················· | ···················· | ···················· | ···················· | ···················· |
| P | 7918 | hCoV-19/USA/CA-CZB-11860/2020\|EPI_ISL_625617\|2020- | ···················· | ·A·················· | ···················· | ···················· | ···················· | ···················· |
| P | 7919 | hCoV-19/USA/CA-CZB-11865/2020\|EPI_ISL_625619\|2020- | ···················· | ·A·················· | ···················· | ···················· | ···················· | ···················· |
| P | 7921 | hCoV-19/USA/CA-CZB-11870/2020\|EPI_ISL_625622\|2020- | ···················· | ···················· | ···················· | ···················· | ···················· | ···················· |
| P | 7922 | hCoV-19/USA/CA-CZB-11968/2020\|EPI_ISL_625623\|2020- | ···················· | ···················· | ···················· | ···················· | ···················· | ···················· |
| P | 7923 | hCoV-19/USA/CA-CZB-11962/2020\|EPI_ISL_625625\|2020- | ···················· | ···················· | ···················· | ···················· | ···················· | ···················· |
| P | 7924 | hCoV-19/USA/CA-CZB-11961/2020\|EPI_ISL_625626\|2020- | ···················· | ···················· | ···················· | ···················· | ···················· | ···················· |
| P | 7925 | hCoV-19/USA/CA-CZB-11783/2020\|EPI_ISL_625627\|2020- | ···················· | ···················· | ···················· | ···················· | ···················· | ···················· |
| P | 7926 | hCoV-19/USA/CA-CZB-11784/2020\|EPI_ISL_625628\|2020- | ···················· | ···················· | ···················· | ···················· | ···················· | ···················· |
| P | 7927 | hCoV-19/USA/CA-CZB-11785/2020\|EPI_ISL_625629\|2020- | ···················· | ···················· | ···················· | ···················· | ···················· | ···················· |
| P | 7928 | hCoV-19/USA/CA-CZB-11786/2020\|EPI_ISL_625630\|2020- | ···················· | ···················· | ···················· | ···················· | ···················· | ···················· |
| P | 7929 | hCoV-19/USA/CA-CZB-11793/2020\|EPI_ISL_625636\|2020- | ···················· | ·A·················· | ···················· | ···················· | ···················· | ···················· |

| | | | | | | | | |
|---|---|---|---|---|---|---|---|---|
| P | 7930 | hCoV-19/USA/CA-CZB-11798/2020\|EPI_ISL_625641\|2020- | ···················· | ·A·················· | ···················· | ···················· | ···················· | ···················· |
| P | 7931 | hCoV-19/USA/CA-CZB-11799/2020\|EPI_ISL_625642\|2020- | ···················· | ···················· | ···················· | ···················· | ···················· | ···················· |
| P | 7932 | hCoV-19/USA/CA-CZB-11800/2020\|EPI_ISL_625643\|2020- | ···················· | ·A·················· | ···················· | ···················· | ···················· | ···················· |
| P | 7933 | hCoV-19/USA/CA-CZB-11801/2020\|EPI_ISL_625644\|2020- | ···················· | ·A·················· | ···················· | ···················· | ···················· | ···················· |
| P | 7936 | hCoV-19/USA/CA-CZB-11806/2020\|EPI_ISL_625648\|2020- | ···················· | ···················· | ···················· | ···················· | ···················· | ···················· |
| P | 7937 | hCoV-19/Denmark/DCGC-776/2020\|EPI_ISL_614880\|2020- | ···················· | ···················· | ···················· | ···················· | ···················· | ···················· |
| P | 7938 | hCoV-19/Denmark/DCGC-787/2020\|EPI_ISL_614881\|2020- | ···················· | ···················· | ···················· | ···················· | ···················· | ···················· |
| P | 7939 | hCoV-19/Denmark/DCGC-788/2020\|EPI_ISL_614882\|2020- | ···················· | ···················· | ···················· | ···················· | ···················· | ···················· |
| P | 7940 | hCoV-19/Denmark/DCGC-792/2020\|EPI_ISL_614883\|2020- | ···················· | ···················· | ···················· | ···················· | ···················· | ···················· |
| P | 7941 | hCoV-19/Denmark/DCGC-949/2020\|EPI_ISL_614884\|2020- | ···················· | ···················· | ···················· | ···················· | ···················· | ···················· |
| P | 7942 | hCoV-19/Denmark/DCGC-920/2020\|EPI_ISL_614885\|2020- | ···················· | ···················· | ···················· | ···················· | ···················· | ···················· |
| P | 7943 | hCoV-19/Denmark/DCGC-929/2020\|EPI_ISL_614886\|2020- | ···················· | ···················· | ···················· | ···················· | ···················· | ···················· |
| P | 7944 | hCoV-19/Denmark/DCGC-932/2020\|EPI_ISL_614887\|2020- | ···················· | ···················· | ···················· | ···················· | ···················· | ···················· |
| P | 7945 | hCoV-19/Denmark/DCGC-933/2020\|EPI_ISL_614888\|2020- | ···················· | ···················· | ···················· | ···················· | ···················· | ···················· |
| P | 7946 | hCoV-19/Italy/SAR-ASSL-08/2020\|EPI_ISL_614889\|2020 | ···················· | ···················· | ···················· | ···················· | ···················· | ···················· |
| P | 7947 | hCoV-19/Switzerland/AG-ETHZ-331066/2020\|EPI_ISL_61 | ···················· | ···················· | ···················· | ···················· | ···················· | ···················· |
| P | 7948 | hCoV-19/Switzerland/AG-ETHZ-331076/2020\|EPI_ISL_61 | ···················· | ···················· | ···················· | ···················· | ···················· | ···················· |
| P | 7949 | hCoV-19/Switzerland/BL-ETHZ-330057/2020\|EPI_ISL_61 | ···················· | ···················· | ···················· | ···················· | ···················· | ···················· |
| P | 7950 | hCoV-19/Switzerland/BL-ETHZ-330061/2020\|EPI_ISL_61 | ···················· | ···················· | ···················· | ···················· | ···················· | ···················· |
| P | 7951 | hCoV-19/Switzerland/BL-ETHZ-330063/2020\|EPI_ISL_61 | ···················· | ···················· | ···················· | ···················· | ······K············· | ···················· |
| P | 7952 | hCoV-19/Switzerland/BL-ETHZ-330064/2020\|EPI_ISL_61 | ···················· | ···················· | ···················· | ···················· | ······K············· | ···················· |
| P | 7953 | hCoV-19/Switzerland/BS-ETHZ-330422/2020\|EPI_ISL_61 | ···················· | ···················· | ···················· | ···················· | ···················· | ···················· |
| P | 7954 | hCoV-19/Switzerland/BE-ETHZ-331448/2020\|EPI_ISL_61 | ···················· | ···················· | ···················· | ···················· | ···················· | ···················· |
| P | 7955 | hCoV-19/Switzerland/BE-ETHZ-331450/2020\|EPI_ISL_61 | ···················· | ···R················ | ···················· | ···················· | ···················· | ···················· |
| P | 7956 | hCoV-19/Switzerland/BE-ETHZ-331452/2020\|EPI_ISL_61 | ···················· | ···················· | ···················· | ···················· | ···················· | ···················· |
| P | 7957 | hCoV-19/Switzerland/BE-ETHZ-332171/2020\|EPI_ISL_61 | ···················· | ···················· | ···················· | ···················· | ···················· | ···················· |
| P | 7958 | hCoV-19/Switzerland/GE-ETHZ-330056/2020\|EPI_ISL_61 | ···················· | ···················· | ···················· | ···················· | ···················· | ···················· |
| P | 7959 | hCoV-19/Switzerland/GE-ETHZ-330065/2020\|EPI_ISL_61 | ···················· | ···················· | ···················· | ···················· | ···················· | ···················· |
| P | 7960 | hCoV-19/Switzerland/GE-ETHZ-331453/2020\|EPI_ISL_61 | ···················· | ···················· | ···················· | ···················· | ···················· | ···················· |
| P | 7961 | hCoV-19/Switzerland/GE-ETHZ-331454/2020\|EPI_ISL_61 | ···················· | ···················· | ···················· | ···················· | ···················· | ···················· |
| P | 7962 | hCoV-19/Switzerland/GE-ETHZ-331455/2020\|EPI_ISL_61 | ···················· | ···················· | ···················· | ···················· | ···················· | ···················· |
| P | 7963 | hCoV-19/Switzerland/GE-ETHZ-331458/2020\|EPI_ISL_61 | ···················· | ···················· | ···················· | ···················· | ···················· | ···················· |
| P | 7964 | hCoV-19/Switzerland/GE-ETHZ-331462/2020\|EPI_ISL_61 | ···················· | ···················· | ···················· | ···················· | ···················· | ···················· |
| P | 7965 | hCoV-19/Switzerland/SO-ETHZ-330511/2020\|EPI_ISL_61 | ···················· | ···················· | ···················· | ···················· | ···················· | ···················· |
| P | 7966 | hCoV-19/Switzerland/SO-ETHZ-331068/2020\|EPI_ISL_61 | ···················· | ···················· | ···················· | ···················· | ···················· | ···················· |
| P | 7967 | hCoV-19/Switzerland/SG-ETHZ-330512/2020\|EPI_ISL_61 | ···················· | ···················· | ···················· | ···················· | ···················· | ···················· |
| P | 7968 | hCoV-19/Switzerland/SG-ETHZ-331457/2020\|EPI_ISL_61 | ···················· | ···················· | ···················· | ···················· | ···················· | ···················· |
| P | 7969 | hCoV-19/Switzerland/SG-ETHZ-331461/2020\|EPI_ISL_61 | ···················· | ···················· | ···················· | ···················· | ···················· | ···················· |
| P | 7970 | hCoV-19/Switzerland/VS-ETHZ-330060/2020\|EPI_ISL_61 | ···················· | ···················· | ···················· | ···················· | ···················· | ···················· |
| P | 7971 | hCoV-19/Switzerland/VS-ETHZ-330062/2020\|EPI_ISL_61 | ···················· | ···················· | ···················· | ···················· | ···················· | ···················· |
| P | 7972 | hCoV-19/Switzerland/VS-ETHZ-330426/2020\|EPI_ISL_61 | ···················· | ···················· | ···················· | ···················· | ···················· | ···················· |
| P | 7973 | hCoV-19/Switzerland/VS-ETHZ-332175/2020\|EPI_ISL_61 | ···················· | ···················· | ···················· | ···················· | ···················· | ···················· |
| P | 7974 | hCoV-19/Switzerland/VD-ETHZ-330054/2020\|EPI_ISL_61 | ···················· | ···················· | ···················· | ···················· | ···················· | ···················· |
| P | 7975 | hCoV-19/Switzerland/VD-ETHZ-330058/2020\|EPI_ISL_61 | ···················· | ···················· | ···················· | ···················· | ···················· | ···················· |
| P | 7976 | hCoV-19/Switzerland/VD-ETHZ-330059/2020\|EPI_ISL_61 | ···················· | ···················· | ···················· | ···················· | ···················· | ···················· |
| P | 7977 | hCoV-19/Switzerland/VD-ETHZ-330248/2020\|EPI_ISL_61 | ···················· | ···················· | ···················· | ···················· | ···················· | ···················· |
| P | 7978 | hCoV-19/Switzerland/VD-ETHZ-330249/2020\|EPI_ISL_61 | ···················· | ···················· | ···················· | ···················· | ···················· | ···················· |
| P | 7979 | hCoV-19/Switzerland/VD-ETHZ-330252/2020\|EPI_ISL_61 | ···················· | ···················· | ···················· | ···················· | ···················· | ···················· |
| P | 7980 | hCoV-19/Denmark/DCGC-7955/2020\|EPI_ISL_617101\|2020 | ···················· | ···················· | ···················· | ···················· | ···················· | ···················· |
| P | 7981 | hCoV-19/Denmark/DCGC-7962/2020\|EPI_ISL_617102\|2020 | ···················· | ···················· | ···················· | ···················· | ···················· | ···················· |
| P | 7982 | hCoV-19/Denmark/DCGC-7925/2020\|EPI_ISL_617103\|2020 | ···················· | ···················· | ···················· | ···················· | ···················· | ···················· |
| P | 7983 | hCoV-19/Denmark/DCGC-7943/2020\|EPI_ISL_617104\|2020 | ···················· | ···················· | ···················· | ···················· | ···················· | ···················· |
| P | 7984 | hCoV-19/Denmark/DCGC-7951/2020\|EPI_ISL_617105\|2020 | ···················· | ···················· | ···················· | ···················· | ···················· | ···················· |
| P | 7985 | hCoV-19/Denmark/DCGC-7957/2020\|EPI_ISL_617106\|2020 | ···················· | ···················· | ···················· | ···················· | ···················· | ···················· |
| P | 7986 | hCoV-19/Denmark/DCGC-1680/2020\|EPI_ISL_617107\|2020 | ···················· | ···················· | ···················· | ···················· | ···················· | ···················· |
| P | 7987 | hCoV-19/Denmark/DCGC-7970/2020\|EPI_ISL_617108\|2020 | ···················· | ···················· | ···················· | ···················· | ···················· | ···················· |
| P | 7988 | hCoV-19/Denmark/DCGC-7983/2020\|EPI_ISL_617109\|2020 | ···················· | ···················· | ···················· | ···················· | ···················· | ···················· |
| P | 7989 | hCoV-19/Denmark/DCGC-2137/2020\|EPI_ISL_618400\|2020 | ···················· | ···················· | ···················· | ···················· | ···················· | ···················· |
| P | 7990 | hCoV-19/Denmark/DCGC-2139/2020\|EPI_ISL_618402\|2020 | ···················· | ···················· | ···················· | ···················· | ···················· | ···················· |
| P | 7991 | hCoV-19/Denmark/DCGC-2140/2020\|EPI_ISL_618403\|2020 | ···················· | ···················· | ···················· | ···················· | ···················· | ···················· |
| P | 7992 | hCoV-19/Denmark/DCGC-2141/2020\|EPI_ISL_618404\|2020 | ···················· | ···················· | ···················· | ···················· | ···················· | ···················· |
| P | 7993 | hCoV-19/Denmark/DCGC-2142/2020\|EPI_ISL_618405\|2020 | ···················· | ···················· | ···················· | ···················· | ···················· | ···················· |
| P | 7994 | hCoV-19/Denmark/DCGC-7520/2020\|EPI_ISL_618406\|2020 | ···················· | ···················· | ···················· | ···················· | ···················· | ···················· |
| P | 7995 | hCoV-19/Denmark/DCGC-7467/2020\|EPI_ISL_618407\|2020 | ···················· | ···················· | ···················· | ···················· | ···················· | ···················· |
| P | 7996 | hCoV-19/Denmark/DCGC-7473/2020\|EPI_ISL_618408\|2020 | ···················· | ···················· | ···················· | ···················· | ···················· | ···················· |
| P | 7997 | hCoV-19/Denmark/DCGC-7491/2020\|EPI_ISL_618410\|2020 | ···················· | ···················· | ···················· | ···················· | ···················· | ···················· |
| P | 7998 | hCoV-19/Denmark/DCGC-7498/2020\|EPI_ISL_618411\|2020 | ···················· | ···················· | ···················· | ···················· | ···················· | ···················· |
| P | 7999 | hCoV-19/Denmark/DCGC-7505/2020\|EPI_ISL_618412\|2020 | ···················· | ···················· | ···················· | ···················· | ···················· | ···················· |
| P | 8000 | hCoV-19/Denmark/DCGC-7513/2020\|EPI_ISL_618414\|2020 | ···················· | ···················· | ···················· | ···················· | ···················· | ···················· |
| P | 8001 | hCoV-19/Denmark/DCGC-7521/2020\|EPI_ISL_618415\|2020 | ···················· | ···················· | ···················· | ···················· | ···················· | ···················· |
| P | 8002 | hCoV-19/Denmark/DCGC-7460/2020\|EPI_ISL_618416\|2020 | ···················· | ···················· | ···················· | ···················· | ···················· | ···················· |
| P | 8003 | hCoV-19/Denmark/DCGC-7468/2020\|EPI_ISL_618417\|2020 | ···················· | ···················· | ···················· | ···················· | ···················· | ···················· |
| P | 8004 | hCoV-19/Denmark/DCGC-7474/2020\|EPI_ISL_618418\|2020 | ···················· | ···················· | ···················· | ···················· | ···················· | ···················· |
| P | 8005 | hCoV-19/Denmark/DCGC-7484/2020\|EPI_ISL_618419\|2020 | ···················· | ···················· | ···················· | ···················· | ···················· | ···················· |
| P | 8006 | hCoV-19/Denmark/DCGC-7499/2020\|EPI_ISL_618421\|2020 | ···················· | ···················· | ···················· | ···················· | ···················· | ···················· |
| P | 8007 | hCoV-19/Denmark/DCGC-7506/2020\|EPI_ISL_618422\|2020 | ···················· | ···················· | ···················· | ···················· | ···················· | ···················· |
| P | 8008 | hCoV-19/Denmark/DCGC-7526/2020\|EPI_ISL_618424\|2020 | ···················· | ·A·················· | ···················· | ···················· | ···················· | ···················· |
| P | 8009 | hCoV-19/Denmark/DCGC-7534/2020\|EPI_ISL_618425\|2020 | ···················· | ···················· | ···················· | ···················· | ···················· | ···················· |
| P | 8010 | hCoV-19/Denmark/DCGC-7546/2020\|EPI_ISL_618427\|2020 | ···················· | ···················· | ···················· | ···················· | ···················· | ···················· |
| P | 8011 | hCoV-19/Denmark/DCGC-7552/2020\|EPI_ISL_618428\|2020 | ···················· | ···················· | ···················· | ···················· | ···················· | ···················· |

| | | | | | | | | |
|---|---|---|---|---|---|---|---|---|
| P | 8012 | hCoV-19/Denmark/DCGC-7565/2020\|EPI_ISL_618429\|2020 | ···················· | ····A··············· | ···················· | ···················· | ···················· | ···················· |
| P | 8013 | hCoV-19/Denmark/DCGC-7571/2020\|EPI_ISL_618430\|2020 | ···················· | ···················· | ···················· | ···················· | ···················· | ···················· |
| P | 8014 | hCoV-19/Denmark/DCGC-7577/2020\|EPI_ISL_618431\|2020 | ···················· | ···················· | ···················· | ···················· | ···················· | ···················· |
| P | 8015 | hCoV-19/Denmark/DCGC-7590/2020\|EPI_ISL_618433\|2020 | ···················· | ···················· | ···················· | ···················· | ···················· | ···················· |
| P | 8016 | hCoV-19/Denmark/DCGC-7527/2020\|EPI_ISL_618434\|2020 | ···················· | ···················· | ···················· | ···················· | ···················· | ···················· |
| P | 8017 | hCoV-19/Denmark/DCGC-7535/2020\|EPI_ISL_618435\|2020 | ···················· | ···················· | ···················· | ···················· | ···················· | ···················· |
| P | 8018 | hCoV-19/Denmark/DCGC-7547/2020\|EPI_ISL_618436\|2020 | ···················· | ···················· | ···················· | ···················· | ···················· | ···················· |
| P | 8019 | hCoV-19/Denmark/DCGC-7553/2020\|EPI_ISL_618437\|2020 | ···················· | ···················· | ···················· | ···················· | ···················· | ···················· |
| P | 8020 | hCoV-19/Denmark/DCGC-7558/2020\|EPI_ISL_618438\|2020 | ···················· | ···················· | ···················· | ···················· | ···················· | ···················· |
| P | 8021 | hCoV-19/Denmark/DCGC-7566/2020\|EPI_ISL_618439\|2020 | ···················· | ···················· | ···················· | ···················· | ···················· | ···················· |
| P | 8022 | hCoV-19/Denmark/DCGC-6885/2020\|EPI_ISL_619730\|2020 | ···················· | ···················· | ···················· | ···················· | ···················· | ···················· |
| P | 8023 | hCoV-19/Denmark/DCGC-6886/2020\|EPI_ISL_619731\|2020 | ···················· | ···················· | ···················· | ···················· | ···················· | ···················· |
| P | 8024 | hCoV-19/Denmark/DCGC-6887/2020\|EPI_ISL_619732\|2020 | ···················· | ···················· | ···················· | ···················· | ···················· | ···················· |
| P | 8025 | hCoV-19/Denmark/DCGC-6888/2020\|EPI_ISL_619733\|2020 | ···················· | ···················· | ···················· | ···················· | ···················· | ···················· |
| P | 8026 | hCoV-19/Denmark/DCGC-6889/2020\|EPI_ISL_619734\|2020 | ···················· | ···················· | ···················· | ···················· | ···················· | ···················· |
| P | 8027 | hCoV-19/Denmark/DCGC-6890/2020\|EPI_ISL_619735\|2020 | ···················· | ···················· | ···················· | ···················· | ···················· | ···················· |
| P | 8028 | hCoV-19/Denmark/DCGC-6894/2020\|EPI_ISL_619739\|2020 | ···················· | ···················· | ···················· | ···················· | ···················· | ···················· |
| P | 8029 | hCoV-19/Denmark/DCGC-6895/2020\|EPI_ISL_619740\|2020 | ···················· | ···················· | ···················· | ···················· | ···················· | ···················· |
| P | 8030 | hCoV-19/Denmark/DCGC-6897/2020\|EPI_ISL_619742\|2020 | ···················· | ···················· | ···················· | ···················· | ···················· | ···················· |
| P | 8031 | hCoV-19/Denmark/DCGC-6899/2020\|EPI_ISL_619744\|2020 | ···················· | ···················· | ···················· | ···················· | ···················· | ···················· |
| P | 8032 | hCoV-19/Denmark/DCGC-6900/2020\|EPI_ISL_619745\|2020 | ···················· | ···················· | ···················· | ···················· | ···················· | ···················· |
| P | 8033 | hCoV-19/Denmark/DCGC-6901/2020\|EPI_ISL_619746\|2020 | ···················· | ···················· | ···················· | ···················· | ···················· | ···················· |
| P | 8034 | hCoV-19/Denmark/DCGC-6902/2020\|EPI_ISL_619747\|2020 | ···················· | ···················· | ···················· | ···················· | ···················· | ···················· |
| P | 8035 | hCoV-19/Denmark/DCGC-6903/2020\|EPI_ISL_619748\|2020 | ···················· | ···················· | ···················· | ···················· | ···················· | ···················· |
| P | 8036 | hCoV-19/Denmark/DCGC-6905/2020\|EPI_ISL_619750\|2020 | ···················· | ···················· | ···················· | ···················· | ···················· | ···················· |
| P | 8037 | hCoV-19/Denmark/DCGC-6906/2020\|EPI_ISL_619752\|2020 | ···················· | ···················· | ···················· | ···················· | ···················· | ···················· |
| P | 8038 | hCoV-19/Denmark/DCGC-6907/2020\|EPI_ISL_619753\|2020 | ···················· | ···················· | ···················· | ···················· | ···················· | ···················· |
| P | 8039 | hCoV-19/Denmark/DCGC-6908/2020\|EPI_ISL_619754\|2020 | ···················· | ···················· | ···················· | ···················· | ···················· | ···················· |
| P | 8040 | hCoV-19/Denmark/DCGC-6909/2020\|EPI_ISL_619755\|2020 | ···················· | ···················· | ···················· | ···················· | ···················· | ···················· |
| P | 8041 | hCoV-19/Denmark/DCGC-6910/2020\|EPI_ISL_619756\|2020 | ···················· | ···················· | ···················· | ···················· | ···················· | ···················· |
| P | 8042 | hCoV-19/Denmark/DCGC-6912/2020\|EPI_ISL_619758\|2020 | ···················· | ···················· | ···················· | ···················· | ···················· | ···················· |
| P | 8043 | hCoV-19/Denmark/DCGC-6913/2020\|EPI_ISL_619759\|2020 | ···················· | ···················· | ···················· | ···················· | ···················· | ···················· |
| P | 8044 | hCoV-19/Denmark/DCGC-6914/2020\|EPI_ISL_619760\|2020 | ···················· | ···················· | ···················· | ···················· | ···················· | ···················· |
| P | 8045 | hCoV-19/Denmark/DCGC-6915/2020\|EPI_ISL_619761\|2020 | ···················· | ···················· | ···················· | ···················· | ···················· | ···················· |
| P | 8046 | hCoV-19/Denmark/DCGC-6917/2020\|EPI_ISL_619763\|2020 | ···················· | ·A·················· | ···················· | ···················· | ···················· | ···A················ |
| P | 8047 | hCoV-19/Denmark/DCGC-6919/2020\|EPI_ISL_619765\|2020 | ···················· | ···················· | ···················· | ···················· | ···················· | ···················· |
| P | 8048 | hCoV-19/Denmark/DCGC-6920/2020\|EPI_ISL_619766\|2020 | ···················· | ···················· | ···················· | ···················· | ···················· | ···················· |
| P | 8049 | hCoV-19/Denmark/DCGC-6921/2020\|EPI_ISL_619767\|2020 | ···················· | ···················· | ···················· | ···················· | ···················· | ···················· |
| P | 8050 | hCoV-19/Denmark/DCGC-6923/2020\|EPI_ISL_619769\|2020 | ···················· | ···················· | ···················· | ···················· | ···················· | ···················· |
| P | 8051 | hCoV-19/England/ALDP-AA12E3/2020\|EPI_ISL_624201\|20 | ···················· | ···················· | ···················· | ···················· | ···················· | ···················· |
| P | 8052 | hCoV-19/England/ALDP-AA2B7E/2020\|EPI_ISL_624202\|20 | ···················· | ···················· | ···················· | ···················· | ···················· | ···················· |
| P | 8053 | hCoV-19/England/ALDP-AA2E84/2020\|EPI_ISL_624203\|20 | ···················· | ···················· | ···················· | ···················· | ···················· | ···················· |
| P | 8054 | hCoV-19/England/ALDP-AA2E1B/2020\|EPI_ISL_624204\|20 | ···················· | ···················· | ···················· | ···················· | ···················· | ···················· |
| P | 8055 | hCoV-19/England/ALDP-AA3DC2/2020\|EPI_ISL_624205\|20 | ···················· | ···················· | ···················· | ···················· | ···················· | ···················· |
| P | 8056 | hCoV-19/Wales/ALDP-AA4909/2020\|EPI_ISL_624206\|2020 | ···················· | ···················· | ···················· | ···················· | ···················· | ···················· |
| P | 8057 | hCoV-19/England/ALDP-AA283B/2020\|EPI_ISL_624207\|20 | ···················· | ···················· | ···················· | ···················· | ···················· | ···················· |
| P | 8058 | hCoV-19/England/ALDP-AA46C6/2020\|EPI_ISL_624208\|20 | ···················· | ···················· | ···················· | ···················· | ···················· | ···················· |
| P | 8059 | hCoV-19/England/ALDP-AA1C7C/2020\|EPI_ISL_624209\|20 | ···················· | ···················· | ···················· | ···················· | ···················· | ···················· |
| P | 8060 | hCoV-19/England/ALDP-AA1FFB/2020\|EPI_ISL_624210\|20 | ···················· | ···················· | ···················· | ···················· | ···················· | ···················· |
| P | 8061 | hCoV-19/England/ALDP-AA1D2E/2020\|EPI_ISL_624211\|20 | ···················· | ···················· | ···················· | ···················· | ···················· | ···················· |
| P | 8062 | hCoV-19/England/ALDP-AA1EC1/2020\|EPI_ISL_624213\|20 | ···················· | ···················· | ···················· | ···················· | ···················· | ···················· |
| P | 8063 | hCoV-19/England/ALDP-AA3A61/2020\|EPI_ISL_624214\|20 | ···················· | ···················· | ···················· | ···················· | ···················· | ···················· |
| P | 8064 | hCoV-19/England/ALDP-AA1553/2020\|EPI_ISL_624215\|20 | ···················· | ···················· | ···················· | ···················· | ···················· | ···················· |
| P | 8065 | hCoV-19/England/ALDP-AA284A/2020\|EPI_ISL_624216\|20 | ···················· | ···················· | ···················· | ···················· | ···················· | ···················· |
| P | 8066 | hCoV-19/England/ALDP-AA4444/2020\|EPI_ISL_624217\|20 | ···················· | ···················· | ···················· | ···················· | ···················· | ···················· |
| P | 8067 | hCoV-19/England/ALDP-AA3B5F/2020\|EPI_ISL_624218\|20 | ···················· | ···················· | ···················· | ···················· | ···················· | ···················· |
| P | 8068 | hCoV-19/England/ALDP-AA2E2A/2020\|EPI_ISL_624219\|20 | ···················· | ···················· | ···················· | ···················· | ···················· | ···················· |
| P | 8069 | hCoV-19/England/ALDP-AA3C89/2020\|EPI_ISL_624220\|20 | ···················· | ···················· | ···················· | ···················· | ···················· | ···················· |
| P | 8070 | hCoV-19/England/ALDP-AA4C2E/2020\|EPI_ISL_624221\|20 | ···················· | ···················· | ···················· | ···················· | ···················· | ···················· |
| P | 8071 | hCoV-19/England/ALDP-AA1E85/2020\|EPI_ISL_624222\|20 | ···················· | ···················· | ···················· | ···················· | ···················· | ···················· |
| P | 8072 | hCoV-19/England/ALDP-AA207F/2020\|EPI_ISL_624223\|20 | ···················· | ···················· | ···················· | ···················· | ···················· | ···················· |
| P | 8073 | hCoV-19/England/ALDP-AA2482/2020\|EPI_ISL_624224\|20 | ···················· | ···················· | ···················· | ···················· | ···················· | ···················· |
| P | 8074 | hCoV-19/England/ALDP-AA2419/2020\|EPI_ISL_624225\|20 | ···················· | ···················· | ···················· | ···················· | ···················· | ···················· |
| P | 8075 | hCoV-19/England/ALDP-AA39FB/2020\|EPI_ISL_624227\|20 | ···················· | ···················· | ···················· | ···················· | ···················· | ···················· |
| P | 8076 | hCoV-19/England/ALDP-AA47E1/2020\|EPI_ISL_624228\|20 | ···················· | ···················· | ···················· | ···················· | ···················· | ···················· |
| P | 8077 | hCoV-19/England/ALDP-AA282C/2020\|EPI_ISL_624229\|20 | ···················· | ···················· | ···················· | ···················· | ···················· | ···················· |
| P | 8078 | hCoV-19/England/ALDP-AA3D68/2020\|EPI_ISL_624230\|20 | ···················· | ···················· | ···················· | ···················· | ···················· | ···················· |
| P | 8079 | hCoV-19/England/ALDP-AA45BA/2020\|EPI_ISL_624231\|20 | ···················· | ···················· | ···················· | ···················· | ···················· | ···················· |
| P | 8080 | hCoV-19/Wales/ALDP-AA22F1/2020\|EPI_ISL_624232\|2020 | ···················· | ···················· | ···················· | ···················· | ···················· | ···················· |
| P | 8081 | hCoV-19/England/ALDP-AA2103/2020\|EPI_ISL_624233\|20 | ···················· | ···················· | ···················· | ···················· | ···················· | ···················· |
| P | 8082 | hCoV-19/England/ALDP-AA4BE5/2020\|EPI_ISL_624234\|20 | ···················· | ···················· | ···················· | ···················· | ···················· | ···················· |
| P | 8083 | hCoV-19/England/ALDP-AA3E92/2020\|EPI_ISL_624235\|20 | ···················· | ···················· | ···················· | ···················· | ···················· | ···················· |
| P | 8084 | hCoV-19/England/ALDP-AA3B31/2020\|EPI_ISL_624236\|20 | ···················· | ···················· | ···················· | ···················· | ···················· | ···················· |
| P | 8085 | hCoV-19/England/ALDP-AA0F83/2020\|EPI_ISL_624237\|20 | ···················· | ···················· | ···················· | ···················· | ···················· | ···················· |
| P | 8086 | hCoV-19/England/ALDP-AA3BD7/2020\|EPI_ISL_624238\|20 | ···················· | ···················· | ···················· | ···················· | ···················· | ···················· |
| P | 8087 | hCoV-19/England/ALDP-AA4787/2020\|EPI_ISL_624239\|20 | ···················· | ··········A········· | ···················· | ···················· | ···················· | ···················· |
| P | 8088 | hCoV-19/England/ALDP-AA23A3/2020\|EPI_ISL_624240\|20 | ···················· | ···················· | ···················· | ···················· | ···················· | ···················· |
| P | 8089 | hCoV-19/England/ALDP-AA47B4/2020\|EPI_ISL_624241\|20 | ···················· | ···················· | ···················· | ···················· | ···················· | ···················· |
| P | 8090 | hCoV-19/England/ALDP-AA1580/2020\|EPI_ISL_624242\|20 | ···················· | ···················· | ···················· | ···················· | ···················· | ···················· |
| P | 8091 | hCoV-19/England/ALDP-AA4B5E/2020\|EPI_ISL_624243\|20 | ···················· | ···················· | ···················· | ···················· | ···················· | ···················· |

| | | | | | | | | |
|---|---|---|---|---|---|---|---|---|
| P | 8092 hCoV-19/England/ALDP-AA1ABE/2020\|EPI_ISL_624245\|20 | ···················· | ···················· | ···················· | ···················· | ···················· | ···················· | |
| P | 8093 hCoV-19/England/ALDP-AA1ED0/2020\|EPI_ISL_624247\|20 | ···················· | ···················· | ···················· | ···················· | ······T············· | ···················· | |
| P | 8094 hCoV-19/England/ALDP-AA1517/2020\|EPI_ISL_624249\|20 | ···················· | ···················· | ···················· | ···················· | ···················· | ···················· | |
| P | 8095 hCoV-19/England/ALDP-AA1FA0/2020\|EPI_ISL_624250\|20 | ···················· | ···················· | ···················· | ···················· | ···················· | ···················· | |
| P | 8096 hCoV-19/England/ALDP-AA4796/2020\|EPI_ISL_624251\|20 | ···················· | ···················· | ···················· | ···················· | ···················· | ···················· | |
| P | 8097 hCoV-19/England/ALDP-AA2859/2020\|EPI_ISL_624253\|20 | ···················· | ···················· | ···················· | ···················· | ···················· | ···················· | |
| P | 8098 hCoV-19/England/ALDP-AA4268/2020\|EPI_ISL_624255\|20 | ···················· | ···················· | ···················· | ···················· | ···················· | ···················· | |
| P | 8099 hCoV-19/England/ALDP-AA4B9A/2020\|EPI_ISL_624256\|20 | ···················· | ···················· | ···················· | ···················· | ···················· | ···················· | |
| P | 8100 hCoV-19/England/ALDP-AA3B04/2020\|EPI_ISL_624257\|20 | ···················· | ···················· | ···················· | ···················· | ···················· | ···················· | |
| P | 8101 hCoV-19/England/ALDP-AA4893/2020\|EPI_ISL_624258\|20 | ···················· | ···················· | ···················· | ···················· | ···················· | ···················· | |
| P | 8102 hCoV-19/England/ALDP-AA230D/2020\|EPI_ISL_624259\|20 | ···················· | ·A·················· | ···················· | ···················· | ···················· | ··A················· | |
| P | 8103 hCoV-19/England/ALDP-AA1F73/2020\|EPI_ISL_624260\|20 | ···················· | ···················· | ···················· | ···················· | ···················· | ···················· | |
| P | 8104 hCoV-19/England/ALDP-AA3D59/2020\|EPI_ISL_624261\|20 | ···················· | ···················· | ···················· | ···················· | ···················· | ···················· | |
| P | 8105 hCoV-19/England/ALDP-AA1F46/2020\|EPI_ISL_624262\|20 | ···················· | ···················· | ···················· | ···················· | ···················· | ···················· | |
| P | 8106 hCoV-19/England/ALDP-AA1D79/2020\|EPI_ISL_624263\|20 | ···················· | ···················· | ···················· | ···················· | ···················· | ···················· | |
| P | 8107 hCoV-19/England/ALDP-AA3A43/2020\|EPI_ISL_624264\|20 | ···················· | ···················· | ···················· | ···················· | ···················· | ···················· | |
| P | 8108 hCoV-19/England/ALDP-AA1016/2020\|EPI_ISL_624265\|20 | ···················· | ···················· | ···················· | ···················· | ···················· | ···················· | |
| P | 8109 hCoV-19/England/ALDP-AA1F82/2020\|EPI_ISL_624267\|20 | ···················· | ···················· | ···················· | ···················· | ···················· | ···················· | |
| P | 8110 hCoV-19/England/ALDP-AA464E/2020\|EPI_ISL_624268\|20 | ···················· | ···················· | ···················· | ···················· | ···················· | ···················· | |
| P | 8111 hCoV-19/England/ALDP-AA15DB/2020\|EPI_ISL_624269\|20 | ···················· | ···················· | ···················· | ···················· | ···················· | ···················· | |
| P | 8112 hCoV-19/England/ALDP-AA2789/2020\|EPI_ISL_624270\|20 | ···················· | ···················· | ···················· | ···················· | ···················· | ···················· | |
| P | 8113 hCoV-19/England/ALDP-AA15CC/2020\|EPI_ISL_624271\|20 | ···················· | ···················· | ···················· | ···················· | ···················· | ···················· | |
| P | 8114 hCoV-19/England/ALDP-AA4480/2020\|EPI_ISL_624272\|20 | ···················· | ···················· | ···················· | ···················· | ···················· | ···················· | |
| P | 8115 hCoV-19/England/ALDP-AA0FDE/2020\|EPI_ISL_624273\|20 | ···················· | ···················· | ···················· | ···················· | ···················· | ···················· | |
| P | 8116 hCoV-19/England/ALDP-AA2DC3/2020\|EPI_ISL_624274\|20 | ···················· | ···················· | ···················· | ···················· | ···················· | ···················· | |
| P | 8117 hCoV-19/England/ALDP-AA15AE/2020\|EPI_ISL_624275\|20 | ···················· | ···················· | ···················· | ···················· | ···················· | ···················· | |
| P | 8118 hCoV-19/England/ALDP-AA10BC/2020\|EPI_ISL_624276\|20 | ···················· | ···················· | ···················· | ···················· | ···················· | ···················· | |
| P | 8119 hCoV-19/England/ALDP-AA3B13/2020\|EPI_ISL_624277\|20 | ···················· | ···················· | ···················· | ···················· | ···················· | ···················· | |
| P | 8120 hCoV-19/England/ALDP-AA3CD4/2020\|EPI_ISL_624278\|20 | ···················· | ···················· | ···················· | ···················· | ···················· | ······G············· | |
| P | 8121 hCoV-19/England/ALDP-AA1C9A/2020\|EPI_ISL_624279\|20 | ···················· | ···················· | ···················· | ···················· | ···················· | ···················· | |
| P | 8122 hCoV-19/USA/CA-CZB-11171/2020\|EPI_ISL_625528\|2020- | ···················· | ·A·················· | ···················· | ···················· | ···················· | ···················· | |
| P | 8123 hCoV-19/USA/CA-CZB-11172/2020\|EPI_ISL_625529\|2020- | ···················· | ·A·················· | ···················· | ···················· | ···················· | ···················· | |
| P | 8125 hCoV-19/USA/CA-CZB-11175/2020\|EPI_ISL_625531\|2020- | ···················· | ·A·················· | ···················· | ···················· | ···················· | ···················· | |
| P | 8127 hCoV-19/USA/CA-CZB-11179/2020\|EPI_ISL_625534\|2020- | ···················· | ·A·················· | ···················· | ···················· | ···················· | ···················· | |
| P | 8128 hCoV-19/USA/CA-CZB-11181/2020\|EPI_ISL_625535\|2020- | ···················· | ·A·················· | ···················· | ···················· | ···················· | ···················· | |
| P | 8130 hCoV-19/USA/CA-CZB-11183/2020\|EPI_ISL_625537\|2020- | ···················· | ·A·················· | ···················· | ···················· | ···················· | ···················· | |
| P | 8131 hCoV-19/USA/CA-CZB-11184/2020\|EPI_ISL_625538\|2020- | ···················· | ·A·················· | ···················· | ···················· | ···················· | ···················· | |
| P | 8132 hCoV-19/USA/CA-CZB-11185/2020\|EPI_ISL_625539\|2020- | ···················· | ·A·················· | ···················· | ···················· | ···················· | ···················· | |
| P | 8133 hCoV-19/USA/CA-CZB-11186/2020\|EPI_ISL_625540\|2020- | ···················· | ·A·················· | ···················· | ···················· | ···················· | ···················· | |
| P | 8135 hCoV-19/USA/CA-CZB-11188/2020\|EPI_ISL_625542\|2020- | ···················· | ·A·················· | ···················· | ···················· | ···················· | ···················· | |
| P | 8138 hCoV-19/USA/CA-CZB-11191/2020\|EPI_ISL_625545\|2020- | ···················· | ·A·················· | ···················· | ···················· | ···················· | ···················· | |
| P | 8140 hCoV-19/USA/CA-CZB-11550/2020\|EPI_ISL_625548\|2020- | ···················· | ···················· | ···················· | ···················· | NNN················· | ···················· | 1 |
| P | 8141 hCoV-19/USA/CA-CZB-11549/2020\|EPI_ISL_625549\|2020- | ···················· | ·A·················· | ···················· | ···················· | ···················· | ···················· | |
| P | 8142 hCoV-19/USA/CA-CZB-11581/2020\|EPI_ISL_625550\|2020- | ···················· | ···················· | ···················· | ···················· | ···················· | ···················· | |
| P | 8143 hCoV-19/USA/CA-CZB-11580/2020\|EPI_ISL_625551\|2020- | ···················· | ···················· | ···················· | ···················· | ···················· | ···················· | |
| P | 8144 hCoV-19/USA/CA-CZB-11579/2020\|EPI_ISL_625552\|2020- | ···················· | ···················· | ···················· | ···················· | ···················· | ···················· | |
| P | 8145 hCoV-19/USA/CA-CZB-11578/2020\|EPI_ISL_625553\|2020- | ···················· | ···················· | ···················· | ···················· | ···················· | ···················· | |
| P | 8146 hCoV-19/USA/CA-CZB-11575/2020\|EPI_ISL_625555\|2020- | ···················· | ···················· | ···················· | ···················· | ···················· | ···················· | |
| P | 8147 hCoV-19/USA/CA-CZB-11574/2020\|EPI_ISL_625556\|2020- | ···················· | ···················· | ···················· | ···················· | ···················· | ···················· | |
| P | 8148 hCoV-19/USA/CA-CZB-11573/2020\|EPI_ISL_625557\|2020- | ···················· | ···················· | ···················· | ···················· | ···················· | ···················· | |
| P | 8149 hCoV-19/USA/CA-CZB-11572/2020\|EPI_ISL_625558\|2020- | ···················· | ···················· | ···················· | ···················· | ···················· | ···················· | |
| P | 8150 hCoV-19/USA/CA-CZB-11589/2020\|EPI_ISL_625559\|2020- | ···················· | ·A·················· | ···················· | ···················· | ···················· | ···················· | |
| P | 8151 hCoV-19/USA/CA-CZB-11588/2020\|EPI_ISL_625560\|2020- | ···················· | ···················· | ···················· | ···················· | ···················· | ···················· | |
| P | 8152 hCoV-19/USA/CA-CZB-11587/2020\|EPI_ISL_625561\|2020- | ···················· | ···················· | ···················· | ···················· | ···················· | ···················· | |
| P | 8153 hCoV-19/USA/CA-CZB-11586/2020\|EPI_ISL_625562\|2020- | ···················· | ···················· | ···················· | ···················· | ···················· | ···················· | |
| P | 8154 hCoV-19/USA/CA-CZB-11585/2020\|EPI_ISL_625563\|2020- | ···················· | ·A·················· | ···················· | ···················· | ···················· | ···················· | |
| P | 8155 hCoV-19/USA/CA-CZB-11584/2020\|EPI_ISL_625564\|2020- | ···················· | ·A·················· | ···················· | ···················· | ···················· | ···················· | |
| P | 8157 hCoV-19/USA/CA-CZB-11582/2020\|EPI_ISL_625566\|2020- | ···················· | ·A·················· | ···················· | ···················· | ···················· | ···················· | |
| P | 8158 hCoV-19/USA/CA-CZB-11547/2020\|EPI_ISL_625567\|2020- | ···················· | ·A·················· | ···················· | ···················· | ···················· | ···················· | |
| P | 8159 hCoV-19/USA/CA-CZB-11546/2020\|EPI_ISL_625568\|2020- | ···················· | ·A·················· | ···················· | ···················· | ···················· | ···················· | |
| P | 8160 hCoV-19/USA/CA-CZB-11544/2020\|EPI_ISL_625569\|2020- | ···················· | ···················· | ···················· | ···················· | ···················· | ···················· | |
| P | 8161 hCoV-19/USA/CA-CZB-11542/2020\|EPI_ISL_625570\|2020- | ···················· | ·A·················· | ···················· | ···················· | ···················· | ···················· | |
| P | 8162 hCoV-19/USA/CA-CZB-11541/2020\|EPI_ISL_625571\|2020- | ···················· | ·A·················· | ···················· | ···················· | ···················· | ···················· | |
| P | 8163 hCoV-19/USA/CA-CZB-11539/2020\|EPI_ISL_625572\|2020- | ···················· | ·A·················· | ···················· | ···················· | ···················· | ···················· | |
| P | 8164 hCoV-19/USA/CA-CZB-11524/2020\|EPI_ISL_625573\|2020- | ···················· | ···················· | ···················· | ···················· | ···················· | ···················· | |
| P | 8167 hCoV-19/USA/CA-CZB-11521/2020\|EPI_ISL_625576\|2020- | ···················· | ·A·················· | ···················· | ···················· | ···················· | ···················· | |
| P | 8168 hCoV-19/USA/CA-CZB-11519/2020\|EPI_ISL_625578\|2020- | ···················· | ·A·················· | ···················· | ···················· | ···················· | ···················· | |
| P | 8169 hCoV-19/USA/CA-CZB-11518/2020\|EPI_ISL_625579\|2020- | ···················· | ·A·················· | ···················· | ···················· | ···················· | ···················· | |
| P | 8170 hCoV-19/USA/CA-CZB-11517/2020\|EPI_ISL_625580\|2020- | ···················· | ·A·················· | ···················· | ···················· | ···················· | ···················· | |
| P | 8172 hCoV-19/USA/CA-CZB-11514/2020\|EPI_ISL_625582\|2020- | ···················· | ·A·················· | ···················· | ···················· | ···················· | ···················· | |
| P | 8173 hCoV-19/USA/CA-CZB-11513/2020\|EPI_ISL_625583\|2020- | ···················· | ·A·················· | ···················· | ···················· | ···················· | ···················· | |
| P | 8174 hCoV-19/USA/CA-CZB-11511/2020\|EPI_ISL_625584\|2020- | ···················· | ·A·················· | ···················· | ···················· | ···················· | ···················· | |
| P | 8175 hCoV-19/USA/CA-CZB-11510/2020\|EPI_ISL_625585\|2020- | ···················· | ·A·················· | ···················· | ···················· | ···················· | ···················· | |
| P | 8176 hCoV-19/USA/CA-CZB-11509/2020\|EPI_ISL_625586\|2020- | ···················· | ·A·················· | ···················· | ···················· | ···················· | ···················· | |
| P | 8177 hCoV-19/USA/CA-CZB-11508/2020\|EPI_ISL_625587\|2020- | ···················· | ·A·················· | ···················· | ···················· | ···················· | ···················· | |
| P | 8178 hCoV-19/USA/CA-CZB-11506/2020\|EPI_ISL_625588\|2020- | ···················· | ···················· | ···················· | ···················· | ···················· | ···················· | |
| P | 8179 hCoV-19/USA/CA-CZB-11505/2020\|EPI_ISL_625589\|2020- | ···················· | ···················· | ···················· | ···················· | ···················· | ···················· | |
| P | 8180 hCoV-19/USA/CA-CZB-11504/2020\|EPI_ISL_625590\|2020- | ···················· | ·A·················· | ···················· | ···················· | ···················· | ···················· | |
| P | 8181 hCoV-19/USA/CA-CZB-11502/2020\|EPI_ISL_625591\|2020- | ···················· | ·A·················· | ···················· | ···················· | ···················· | ···················· | |
| P | 8182 hCoV-19/USA/CA-CZB-11501/2020\|EPI_ISL_625592\|2020- | ···················· | ·A·················· | ···················· | ···················· | ···················· | ···················· | |

| | | | | | | | |
|---|---|---|---|---|---|---|---|
| P | 8183 hCoV-19/USA/CA-CZB-11872/2020\|EPI_ISL_625594\|2020- | ···················· | ···················· | ···················· | ···················· | ···················· | ···················· |
| P | 8184 hCoV-19/USA/CA-CZB-11873/2020\|EPI_ISL_625595\|2020- | ···················· | ···················· | ···················· | ···················· | ···················· | ···················· |
| P | 8185 hCoV-19/USA/CA-CZB-11874/2020\|EPI_ISL_625596\|2020- | ···················· | ···················· | ···················· | ···················· | ···················· | ···················· |
| P | 8186 hCoV-19/USA/CA-CZB-11875/2020\|EPI_ISL_625597\|2020- | ···················· | ···················· | ···················· | ···················· | ···················· | ···················· |
| P | 8187 hCoV-19/USA/CA-CZB-11876/2020\|EPI_ISL_625598\|2020- | ···················· | ···················· | ···················· | ···················· | ···················· | ···················· |
| P | 8188 hCoV-19/USA/CA-CZB-11879/2020\|EPI_ISL_625599\|2020- | ···················· | ···················· | ···················· | ···················· | ···················· | ···················· |
| P | 8189 hCoV-19/USA/CA-CZB-11908/2020\|EPI_ISL_625600\|2020- | ···················· | ···················· | ···················· | ···················· | ···················· | ···················· |
| P | 8190 hCoV-19/USA/CA-CZB-11909/2020\|EPI_ISL_625601\|2020- | ···················· | ···················· | ···················· | ···················· | ···················· | ···················· |
| P | 8191 hCoV-19/USA/CA-CZB-11920/2020\|EPI_ISL_625602\|2020- | ···················· | ··C················· | ···················· | ···················· | ···················· | ···················· |
| P | 8193 hCoV-19/Denmark/DCGC-1190/2020\|EPI_ISL_614836\|2020 | ···················· | ···················· | ···················· | ···················· | ···················· | ···················· |
| P | 8194 hCoV-19/Denmark/DCGC-1191/2020\|EPI_ISL_614837\|2020 | ···················· | ···················· | ···················· | ···················· | ···················· | ···················· |
| P | 8195 hCoV-19/Denmark/DCGC-1192/2020\|EPI_ISL_614838\|2020 | ···················· | ···················· | ···················· | ···················· | ···················· | ···················· |
| P | 8196 hCoV-19/Denmark/DCGC-1193/2020\|EPI_ISL_614839\|2020 | ···················· | ···················· | ···················· | ···················· | ···················· | ···················· |
| P | 8197 hCoV-19/Denmark/DCGC-1194/2020\|EPI_ISL_614840\|2020 | ···················· | ···················· | ···················· | ···················· | ···················· | ···················· |
| P | 8198 hCoV-19/Denmark/DCGC-1195/2020\|EPI_ISL_614841\|2020 | ···················· | ···················· | ···················· | ···················· | ···················· | ···················· |
| P | 8199 hCoV-19/Denmark/DCGC-1196/2020\|EPI_ISL_614842\|2020 | ···················· | ···················· | ···················· | ···················· | ···················· | ···················· |
| P | 8200 hCoV-19/Denmark/DCGC-1197/2020\|EPI_ISL_614843\|2020 | ···················· | ···················· | ···················· | ···················· | ···················· | ···················· |
| P | 8201 hCoV-19/Denmark/DCGC-1198/2020\|EPI_ISL_614844\|2020 | ···················· | ···················· | ···················· | ···················· | ···················· | ···················· |
| P | 8202 hCoV-19/Denmark/DCGC-1199/2020\|EPI_ISL_614845\|2020 | ···················· | ···················· | ···················· | ···················· | ···················· | ···················· |
| P | 8203 hCoV-19/Denmark/DCGC-1200/2020\|EPI_ISL_614846\|2020 | ···················· | ···················· | ···················· | ···················· | ···················· | ···················· |
| P | 8204 hCoV-19/Denmark/DCGC-1205/2020\|EPI_ISL_614850\|2020 | ···················· | ···················· | ···················· | ···················· | ···················· | ···················· |
| P | 8205 hCoV-19/Denmark/DCGC-1206/2020\|EPI_ISL_614851\|2020 | ···················· | ···················· | ···················· | ···················· | ···················· | ···················· |
| P | 8206 hCoV-19/Denmark/DCGC-1207/2020\|EPI_ISL_614852\|2020 | ···················· | ···················· | ···················· | ···················· | ···················· | ···················· |
| P | 8207 hCoV-19/Denmark/DCGC-1208/2020\|EPI_ISL_614853\|2020 | ···················· | ···················· | ···················· | ···················· | ···················· | ···················· |
| P | 8208 hCoV-19/Denmark/DCGC-1213/2020\|EPI_ISL_614856\|2020 | ···················· | ···················· | ···················· | ···················· | ···················· | ···················· |
| P | 8209 hCoV-19/Denmark/DCGC-750/2020\|EPI_ISL_614857\|2020- | ···················· | ···················· | ···················· | ···················· | ···················· | ···················· |
| P | 8210 hCoV-19/Denmark/DCGC-8056/2020\|EPI_ISL_617000\|2020 | ···················· | ······A············· | ···················· | ···················· | ···················· | ···················· |
| P | 8211 hCoV-19/Denmark/DCGC-8117/2020\|EPI_ISL_617001\|2020 | ···················· | ···················· | ···················· | ···················· | ···················· | ···················· |
| P | 8212 hCoV-19/Denmark/DCGC-8161/2020\|EPI_ISL_617002\|2020 | ···················· | ···················· | ···················· | ···················· | ···················· | ···················· |
| P | 8213 hCoV-19/Denmark/DCGC-8186/2020\|EPI_ISL_617003\|2020 | ···················· | ···················· | ···················· | ···················· | ···················· | ···················· |
| P | 8214 hCoV-19/Denmark/DCGC-8132/2020\|EPI_ISL_617004\|2020 | ···················· | ···················· | ···················· | ···················· | ···················· | ···················· |
| P | 8215 hCoV-19/Denmark/DCGC-8187/2020\|EPI_ISL_617005\|2020 | ···················· | ···················· | ···················· | ···················· | ···················· | ···················· |
| P | 8216 hCoV-19/Denmark/DCGC-8119/2020\|EPI_ISL_617006\|2020 | ···················· | ···················· | ···················· | ···················· | ···················· | ···················· |
| P | 8217 hCoV-19/Denmark/DCGC-8148/2020\|EPI_ISL_617007\|2020 | ···················· | ···················· | ···················· | ···················· | ···················· | ···················· |
| P | 8218 hCoV-19/Denmark/DCGC-8122/2020\|EPI_ISL_617008\|2020 | ···················· | ···················· | ···················· | ···················· | ···················· | ···················· |
| P | 8219 hCoV-19/Denmark/DCGC-8164/2020\|EPI_ISL_617009\|2020 | ···················· | ···················· | ···················· | ···················· | ···················· | ···················· |
| P | 8220 hCoV-19/Denmark/DCGC-8129/2020\|EPI_ISL_617010\|2020 | ···················· | ···················· | ···················· | ···················· | ···················· | ···················· |
| P | 8221 hCoV-19/Denmark/DCGC-8170/2020\|EPI_ISL_617011\|2020 | ···················· | ···················· | ···················· | ···················· | ···················· | ···················· |
| P | 8222 hCoV-19/Denmark/DCGC-8184/2020\|EPI_ISL_617012\|2020 | ···················· | ···················· | ···················· | ···················· | ···················· | ···················· |
| P | 8223 hCoV-19/Denmark/DCGC-8178/2020\|EPI_ISL_617013\|2020 | ···················· | ···················· | ···················· | ···················· | ···················· | ···················· |
| P | 8224 hCoV-19/Denmark/DCGC-8219/2020\|EPI_ISL_617014\|2020 | ···················· | ···················· | ···················· | ···················· | ···················· | ···················· |
| P | 8225 hCoV-19/Denmark/DCGC-8245/2020\|EPI_ISL_617015\|2020 | ···················· | ···················· | ···················· | ···················· | ···················· | ···················· |
| P | 8226 hCoV-19/Denmark/DCGC-8255/2020\|EPI_ISL_617016\|2020 | ···················· | ···················· | ···················· | ···················· | ···················· | ···················· |
| P | 8227 hCoV-19/Denmark/DCGC-8191/2020\|EPI_ISL_617017\|2020 | ···················· | ···················· | ···················· | ···················· | ···················· | ···················· |
| P | 8228 hCoV-19/Denmark/DCGC-8192/2020\|EPI_ISL_617018\|2020 | ···················· | ···················· | ···················· | ···················· | ···················· | ···················· |
| P | 8229 hCoV-19/Denmark/DCGC-8205/2020\|EPI_ISL_617019\|2020 | ···················· | ···················· | ···················· | ···················· | ···················· | ···················· |
| P | 8230 hCoV-19/Denmark/DCGC-8211/2020\|EPI_ISL_617020\|2020 | ···················· | ···················· | ···················· | ···················· | ···················· | ···················· |
| P | 8231 hCoV-19/Denmark/DCGC-8242/2020\|EPI_ISL_617021\|2020 | ···················· | ···················· | ···················· | ···················· | ···················· | ···················· |
| P | 8232 hCoV-19/Denmark/DCGC-8252/2020\|EPI_ISL_617022\|2020 | ···················· | ···················· | ···················· | ···················· | ···················· | ···················· |
| P | 8233 hCoV-19/Denmark/DCGC-8206/2020\|EPI_ISL_617023\|2020 | ···················· | ···················· | ···················· | ···················· | ···················· | ···················· |
| P | 8234 hCoV-19/Denmark/DCGC-8243/2020\|EPI_ISL_617024\|2020 | ···················· | ···················· | ···················· | ···················· | ···················· | ···················· |
| P | 8235 hCoV-19/Denmark/DCGC-8247/2020\|EPI_ISL_617025\|2020 | ···················· | ···················· | ···················· | ···················· | ···················· | ···················· |
| P | 8236 hCoV-19/Denmark/DCGC-8208/2020\|EPI_ISL_617026\|2020 | ···················· | ···················· | ···················· | ···················· | ···················· | ···················· |
| P | 8237 hCoV-19/Denmark/DCGC-8281/2020\|EPI_ISL_617027\|2020 | ···················· | ···················· | ···················· | ···················· | ···················· | ···················· |
| P | 8238 hCoV-19/Denmark/DCGC-8269/2020\|EPI_ISL_617028\|2020 | ···················· | ···················· | ···················· | ···················· | ···················· | ···················· |
| P | 8239 hCoV-19/Denmark/DCGC-8325/2020\|EPI_ISL_617029\|2020 | ···················· | ···················· | ···················· | ···················· | ···················· | ···················· |
| P | 8240 hCoV-19/Denmark/DCGC-8270/2020\|EPI_ISL_617030\|2020 | ···················· | ···················· | ···················· | ···················· | ···················· | ···················· |
| P | 8241 hCoV-19/Denmark/DCGC-8286/2020\|EPI_ISL_617031\|2020 | ···················· | ···················· | ···················· | ···················· | ···················· | ···················· |
| P | 8242 hCoV-19/Denmark/DCGC-8302/2020\|EPI_ISL_617032\|2020 | ···················· | ···················· | ···················· | ···················· | ···················· | ···················· |
| P | 8243 hCoV-19/Denmark/DCGC-8277/2020\|EPI_ISL_617033\|2020 | ···················· | ···················· | ···················· | ···················· | ···················· | ···················· |
| P | 8244 hCoV-19/Denmark/DCGC-8353/2020\|EPI_ISL_617034\|2020 | ···················· | ···················· | ···················· | ···················· | ···················· | ···················· |
| P | 8245 hCoV-19/Denmark/DCGC-8402/2020\|EPI_ISL_617035\|2020 | ···················· | ···················· | ···················· | ···················· | ···················· | ···················· |
| P | 8246 hCoV-19/Denmark/DCGC-8355/2020\|EPI_ISL_617036\|2020 | ···················· | ···················· | ···················· | ···················· | ···················· | ···················· |
| P | 8247 hCoV-19/Denmark/DCGC-8382/2020\|EPI_ISL_617037\|2020 | ···················· | ···················· | ···················· | ···················· | ···················· | ···················· |
| P | 8248 hCoV-19/Denmark/DCGC-8362/2020\|EPI_ISL_617038\|2020 | ···················· | ···················· | ···················· | ···················· | ···················· | ···················· |
| P | 8249 hCoV-19/Denmark/DCGC-8374/2020\|EPI_ISL_617039\|2020 | ···················· | ···················· | ···················· | ···················· | ···················· | ···················· |
| P | 8250 hCoV-19/Denmark/DCGC-8391/2020\|EPI_ISL_617040\|2020 | ···················· | ···················· | ···················· | ···················· | ···················· | ···················· |
| P | 8251 hCoV-19/Denmark/DCGC-8357/2020\|EPI_ISL_617041\|2020 | ···················· | ···················· | ···················· | ···················· | ···················· | ···················· |
| P | 8252 hCoV-19/Denmark/DCGC-8367/2020\|EPI_ISL_617042\|2020 | ···················· | ···················· | ···················· | ···················· | ···················· | ···················· |
| P | 8253 hCoV-19/Denmark/DCGC-8428/2020\|EPI_ISL_617043\|2020 | ···················· | ···················· | ···················· | ···················· | ···················· | ···················· |
| P | 8254 hCoV-19/Denmark/DCGC-8434/2020\|EPI_ISL_617044\|2020 | ···················· | ···················· | ···················· | ···················· | ···················· | ···················· |
| P | 8255 hCoV-19/Denmark/DCGC-8423/2020\|EPI_ISL_617045\|2020 | ···················· | ···················· | ···················· | ···················· | ···················· | ···················· |
| P | 8256 hCoV-19/Denmark/DCGC-8435/2020\|EPI_ISL_617046\|2020 | ···················· | ···················· | ···················· | ···················· | ···················· | ···················· |
| P | 8257 hCoV-19/Denmark/DCGC-8049/2020\|EPI_ISL_617047\|2020 | ···················· | ·A·················· | ···················· | ···················· | ···················· | ···A················ |
| P | 8258 hCoV-19/Denmark/DCGC-8221/2020\|EPI_ISL_617048\|2020 | ···················· | ·A·················· | ···················· | ···················· | ···················· | ···A················ |
| P | 8259 hCoV-19/Denmark/DCGC-8222/2020\|EPI_ISL_617049\|2020 | ···················· | ·A·················· | ···················· | ···················· | ···················· | ···A················ |
| P | 8260 hCoV-19/Denmark/DCGC-8289/2020\|EPI_ISL_617050\|2020 | ···················· | ·A·················· | ···················· | ···················· | ···················· | ···A················ |
| P | 8261 hCoV-19/Denmark/DCGC-8297/2020\|EPI_ISL_617051\|2020 | ···················· | ·A·················· | ···················· | ···················· | ···················· | ···A················ |
| P | 8262 hCoV-19/Denmark/DCGC-8364/2020\|EPI_ISL_617052\|2020 | ···················· | ·A·················· | ···················· | ···················· | ···················· | ···A················ |
| P | 8263 hCoV-19/Denmark/DCGC-8365/2020\|EPI_ISL_617053\|2020 | ···················· | ·A·················· | ···················· | ···················· | ···················· | ···A················ |

| | | | | | | | |
|---|---|---|---|---|---|---|---|
| P | 8264 hCoV-19/Denmark/DCGC-8375/2020\|EPI_ISL_617054\|2020 | ···················· | ·A·················· | ···················· | ···················· | ···················· | ···A················ |
| P | 8265 hCoV-19/Denmark/DCGC-7939/2020\|EPI_ISL_617100\|2020 | ···················· | ·A·················· | ···················· | ···················· | ···················· | ···················· |
| P | 8266 hCoV-19/Denmark/DCGC-1989/2020\|EPI_ISL_618320\|2020 | ···················· | ···················· | ···················· | ···················· | ···················· | ···················· |
| P | 8267 hCoV-19/Denmark/DCGC-2067/2020\|EPI_ISL_618321\|2020 | ···················· | ···················· | ···················· | ···················· | ···················· | ···················· |
| P | 8268 hCoV-19/Denmark/DCGC-2068/2020\|EPI_ISL_618322\|2020 | ···················· | ···················· | ···················· | ···················· | ···················· | ···················· |
| P | 8269 hCoV-19/Denmark/DCGC-1980/2020\|EPI_ISL_618323\|2020 | ···················· | ···················· | ···················· | ···················· | ···················· | ···················· |
| P | 8270 hCoV-19/Denmark/DCGC-1981/2020\|EPI_ISL_618324\|2020 | ···················· | ···················· | ···················· | ···················· | ···················· | ···················· |
| P | 8271 hCoV-19/Denmark/DCGC-1982/2020\|EPI_ISL_618325\|2020 | ···················· | ···················· | ···················· | ···················· | ···················· | ···················· |
| P | 8272 hCoV-19/Denmark/DCGC-2035/2020\|EPI_ISL_618326\|2020 | ···················· | ···················· | ···················· | ···················· | ···················· | ···················· |
| P | 8273 hCoV-19/Denmark/DCGC-1953/2020\|EPI_ISL_618327\|2020 | ···················· | ·A·················· | ···················· | ···················· | ···················· | ···A················ |
| P | 8274 hCoV-19/Denmark/DCGC-1954/2020\|EPI_ISL_618328\|2020 | ···················· | ···················· | ···················· | ···················· | ···················· | ···················· |
| P | 8275 hCoV-19/Denmark/DCGC-1983/2020\|EPI_ISL_618329\|2020 | ···················· | ···················· | ···················· | ···················· | ···················· | ···················· |
| P | 8276 hCoV-19/Denmark/DCGC-1990/2020\|EPI_ISL_618331\|2020 | ···················· | ···················· | ···················· | ···················· | ···················· | ···················· |
| P | 8277 hCoV-19/Denmark/DCGC-1998/2020\|EPI_ISL_618333\|2020 | ···················· | ···················· | ···················· | ···················· | ···················· | ···················· |
| P | 8278 hCoV-19/Denmark/DCGC-1999/2020\|EPI_ISL_618334\|2020 | ···················· | ···················· | ···················· | ···················· | ···················· | ···················· |
| P | 8279 hCoV-19/Denmark/DCGC-2000/2020\|EPI_ISL_618335\|2020 | ···················· | ···················· | ···················· | ···················· | ···················· | ···················· |
| P | 8280 hCoV-19/Denmark/DCGC-2001/2020\|EPI_ISL_618336\|2020 | ···················· | ···················· | ···················· | ···················· | ···················· | ···················· |
| P | 8281 hCoV-19/Denmark/DCGC-2081/2020\|EPI_ISL_618337\|2020 | ···················· | ···················· | ···················· | ···················· | ···················· | ···················· |
| P | 8282 hCoV-19/Denmark/DCGC-2082/2020\|EPI_ISL_618338\|2020 | ···················· | ···················· | ···················· | ···················· | ···················· | ···················· |
| P | 8283 hCoV-19/Denmark/DCGC-2083/2020\|EPI_ISL_618339\|2020 | ···················· | ···················· | ···················· | ···················· | ···················· | ···················· |
| P | 8284 hCoV-19/Denmark/DCGC-2084/2020\|EPI_ISL_618340\|2020 | ···················· | ···················· | ···················· | ···················· | ···················· | ···················· |
| P | 8285 hCoV-19/Denmark/DCGC-2085/2020\|EPI_ISL_618341\|2020 | ···················· | ···················· | ···················· | ···················· | ···················· | ···················· |
| P | 8286 hCoV-19/Denmark/DCGC-1709/2020\|EPI_ISL_618342\|2020 | ···················· | ···················· | ···················· | ···················· | ···················· | ···················· |
| P | 8287 hCoV-19/Denmark/DCGC-2086/2020\|EPI_ISL_618343\|2020 | ···················· | ···················· | ···················· | ···················· | ···················· | ···················· |
| P | 8288 hCoV-19/Denmark/DCGC-2087/2020\|EPI_ISL_618344\|2020 | ···················· | ···················· | ···················· | ···················· | ···················· | ···················· |
| P | 8289 hCoV-19/Denmark/DCGC-2006/2020\|EPI_ISL_618345\|2020 | ···················· | ···················· | ···················· | ···················· | ···················· | ···················· |
| P | 8290 hCoV-19/Denmark/DCGC-2104/2020\|EPI_ISL_618346\|2020 | ···················· | ···················· | ···················· | ···················· | ···················· | ···················· |
| P | 8291 hCoV-19/Denmark/DCGC-2088/2020\|EPI_ISL_618347\|2020 | ···················· | ···················· | ···················· | ···················· | ···················· | ···················· |
| P | 8292 hCoV-19/Denmark/DCGC-2089/2020\|EPI_ISL_618348\|2020 | ···················· | ···················· | ···················· | ···················· | ···················· | ···················· |
| P | 8293 hCoV-19/Denmark/DCGC-2091/2020\|EPI_ISL_618350\|2020 | ···················· | ···················· | ···················· | ···················· | ···················· | ···················· |
| P | 8294 hCoV-19/Denmark/DCGC-2092/2020\|EPI_ISL_618351\|2020 | ···················· | ···················· | ···················· | ···················· | ···················· | ···················· |
| P | 8295 hCoV-19/Denmark/DCGC-2093/2020\|EPI_ISL_618352\|2020 | ···················· | ···················· | ···················· | ···················· | ···················· | ···················· |
| P | 8296 hCoV-19/Denmark/DCGC-2094/2020\|EPI_ISL_618353\|2020 | ···················· | ···················· | ···················· | ···················· | ···················· | ···················· |
| P | 8297 hCoV-19/Denmark/DCGC-2095/2020\|EPI_ISL_618354\|2020 | ···················· | ···················· | ···················· | ···················· | ···················· | ···················· |
| P | 8298 hCoV-19/Denmark/DCGC-2096/2020\|EPI_ISL_618355\|2020 | ···················· | ···················· | ···················· | ···················· | ···················· | ···················· |
| P | 8299 hCoV-19/Denmark/DCGC-2097/2020\|EPI_ISL_618356\|2020 | ···················· | ···················· | ···················· | ···················· | ···················· | ···················· |
| P | 8300 hCoV-19/Denmark/DCGC-2098/2020\|EPI_ISL_618357\|2020 | ···················· | ···················· | ···················· | ···················· | ···················· | ···················· |
| P | 8301 hCoV-19/Denmark/DCGC-2099/2020\|EPI_ISL_618358\|2020 | ···················· | ···················· | ···················· | ···················· | ···················· | ···················· |
| P | 8302 hCoV-19/Denmark/DCGC-2100/2020\|EPI_ISL_618359\|2020 | ···················· | ···················· | ···················· | ···················· | ···················· | ···················· |
| P | 8303 hCoV-19/Denmark/DCGC-2101/2020\|EPI_ISL_618360\|2020 | ···················· | ···················· | ···················· | ···················· | ···················· | ···················· |
| P | 8304 hCoV-19/Denmark/DCGC-2102/2020\|EPI_ISL_618361\|2020 | ···················· | ···················· | ···················· | ···················· | ···················· | ···················· |
| P | 8305 hCoV-19/Denmark/DCGC-2471/2020\|EPI_ISL_618363\|2020 | ···················· | ···················· | ···················· | ···················· | ···················· | ···················· |
| P | 8306 hCoV-19/Denmark/DCGC-2443/2020\|EPI_ISL_618364\|2020 | ···················· | ···················· | ···················· | ···················· | ···················· | ···················· |
| P | 8307 hCoV-19/Denmark/DCGC-2444/2020\|EPI_ISL_618365\|2020 | ···················· | ···················· | ···················· | ···················· | ···················· | ···················· |
| P | 8308 hCoV-19/Denmark/DCGC-2445/2020\|EPI_ISL_618366\|2020 | ···················· | ···················· | ···················· | ···················· | ···················· | ···················· |
| P | 8309 hCoV-19/Denmark/DCGC-2105/2020\|EPI_ISL_618367\|2020 | ···················· | ···················· | ···················· | ···················· | ···················· | ···················· |
| P | 8310 hCoV-19/Denmark/DCGC-2106/2020\|EPI_ISL_618368\|2020 | ···················· | ···················· | ···················· | ···················· | ···················· | ···················· |
| P | 8311 hCoV-19/Denmark/DCGC-2107/2020\|EPI_ISL_618369\|2020 | ···················· | ···················· | ···················· | ···················· | ···················· | ···················· |
| P | 8312 hCoV-19/Denmark/DCGC-2108/2020\|EPI_ISL_618370\|2020 | ···················· | ···················· | ···················· | ···················· | ···················· | ···················· |
| P | 8313 hCoV-19/Denmark/DCGC-2109/2020\|EPI_ISL_618371\|2020 | ···················· | ···················· | ···················· | ···················· | ···················· | ···················· |
| P | 8314 hCoV-19/Denmark/DCGC-2110/2020\|EPI_ISL_618372\|2020 | ···················· | ···················· | ···················· | ···················· | ···················· | ···················· |
| P | 8315 hCoV-19/Denmark/DCGC-2112/2020\|EPI_ISL_618374\|2020 | ···················· | ···················· | ···················· | ···················· | ···················· | ···················· |
| P | 8316 hCoV-19/Denmark/DCGC-2113/2020\|EPI_ISL_618375\|2020 | ···················· | ···················· | ···················· | ···················· | ···················· | ···················· |
| P | 8317 hCoV-19/Denmark/DCGC-2114/2020\|EPI_ISL_618376\|2020 | ···················· | ···················· | ···················· | ···················· | ···················· | ···················· |
| P | 8318 hCoV-19/Denmark/DCGC-2115/2020\|EPI_ISL_618377\|2020 | ···················· | ···················· | ···················· | ···················· | ···················· | ···················· |
| P | 8319 hCoV-19/Denmark/DCGC-2116/2020\|EPI_ISL_618378\|2020 | ···················· | ···················· | ···················· | ···················· | ···················· | ···················· |
| P | 8320 hCoV-19/Denmark/DCGC-2117/2020\|EPI_ISL_618379\|2020 | ···················· | ···················· | ···················· | ···················· | ···················· | ···················· |
| P | 8321 hCoV-19/Denmark/DCGC-2118/2020\|EPI_ISL_618380\|2020 | ···················· | ···················· | ···················· | ···················· | ···················· | ···················· |
| P | 8322 hCoV-19/Denmark/DCGC-2119/2020\|EPI_ISL_618381\|2020 | ···················· | ···················· | ···················· | ···················· | ···················· | ···················· |
| P | 8324 hCoV-19/Denmark/DCGC-2120/2020\|EPI_ISL_618383\|2020 | ···················· | ···················· | ···················· | ···················· | ···················· | ···················· |
| P | 8325 hCoV-19/Denmark/DCGC-2121/2020\|EPI_ISL_618384\|2020 | ···················· | ···················· | ···················· | ···················· | ···················· | ···················· |
| P | 8326 hCoV-19/Denmark/DCGC-2122/2020\|EPI_ISL_618385\|2020 | ···················· | ···················· | ···················· | ···················· | ···················· | ···················· |
| P | 8327 hCoV-19/Denmark/DCGC-2123/2020\|EPI_ISL_618386\|2020 | ···················· | ···················· | ···················· | ···················· | ···················· | ···················· |
| P | 8328 hCoV-19/Denmark/DCGC-2124/2020\|EPI_ISL_618387\|2020 | ···················· | ···················· | ···················· | ···················· | ···················· | ···················· |
| P | 8329 hCoV-19/Denmark/DCGC-2126/2020\|EPI_ISL_618389\|2020 | ···················· | ···················· | ···················· | ···················· | ···················· | ···················· |
| P | 8330 hCoV-19/Denmark/DCGC-2127/2020\|EPI_ISL_618390\|2020 | ···················· | ···················· | ···················· | ···················· | ···················· | ···················· |
| P | 8331 hCoV-19/Denmark/DCGC-2128/2020\|EPI_ISL_618391\|2020 | ···················· | ···················· | ···················· | ···················· | ···················· | ···················· |
| P | 8332 hCoV-19/Denmark/DCGC-2129/2020\|EPI_ISL_618392\|2020 | ···················· | ···················· | ···················· | ···················· | ···················· | ···················· |
| P | 8333 hCoV-19/Denmark/DCGC-2130/2020\|EPI_ISL_618393\|2020 | ···················· | ···················· | ···················· | ···················· | ···················· | ···················· |
| P | 8334 hCoV-19/Denmark/DCGC-2131/2020\|EPI_ISL_618394\|2020 | ···················· | ···················· | ···················· | ···················· | ···················· | ···················· |
| P | 8335 hCoV-19/Denmark/DCGC-2132/2020\|EPI_ISL_618395\|2020 | ···················· | ···················· | ···················· | ···················· | ···················· | ···················· |
| P | 8336 hCoV-19/Denmark/DCGC-2133/2020\|EPI_ISL_618396\|2020 | ···················· | ···················· | ···················· | ···················· | ···················· | ···················· |
| P | 8337 hCoV-19/Denmark/DCGC-2134/2020\|EPI_ISL_618397\|2020 | ···················· | ···················· | ···················· | ···················· | ···················· | ···················· |
| P | 8338 hCoV-19/Denmark/DCGC-2135/2020\|EPI_ISL_618398\|2020 | ···················· | ···················· | ···················· | ···················· | ···················· | ···················· |
| P | 8339 hCoV-19/Denmark/DCGC-2136/2020\|EPI_ISL_618399\|2020 | ···················· | ···················· | ···················· | ···················· | ···················· | ···················· |
| P | 8340 hCoV-19/Denmark/DCGC-6602/2020\|EPI_ISL_619650\|2020 | ···················· | ···················· | ···················· | ···················· | ···················· | ···················· |
| P | 8341 hCoV-19/Denmark/DCGC-6604/2020\|EPI_ISL_619652\|2020 | ···················· | ···················· | ···················· | ···················· | ···················· | ···················· |
| P | 8342 hCoV-19/Denmark/DCGC-6606/2020\|EPI_ISL_619654\|2020 | ···················· | ···················· | ···················· | ···················· | ···················· | ···················· |
| P | 8343 hCoV-19/Denmark/DCGC-6607/2020\|EPI_ISL_619655\|2020 | ···················· | ···················· | ···················· | ···················· | ···················· | ···················· |
| P | 8344 hCoV-19/Denmark/DCGC-6611/2020\|EPI_ISL_619660\|2020 | ···················· | ···················· | ···················· | ···················· | ···················· | ···················· |

| | | | | | | | | |
|---|---|---|---|---|---|---|---|---|
| P | 8345 | hCoV-19/Denmark/DCGC-6613/2020\|EPI_ISL_619662\|2020 | ···················· | ···················· | ····················· | ···················· | ···················· | ···················· |
| P | 8346 | hCoV-19/Denmark/DCGC-6614/2020\|EPI_ISL_619663\|2020 | ···················· | ···················· | ····················· | ···················· | ···················· | ···················· |
| P | 8347 | hCoV-19/Denmark/DCGC-6615/2020\|EPI_ISL_619664\|2020 | ···················· | ···················· | ····················· | ···················· | ···················· | ···················· |
| P | 8348 | hCoV-19/Denmark/DCGC-6616/2020\|EPI_ISL_619665\|2020 | ···················· | ···················· | ····················· | ···················· | ···················· | ···················· |
| P | 8349 | hCoV-19/Denmark/DCGC-6617/2020\|EPI_ISL_619666\|2020 | ···················· | ···················· | ····················· | ···················· | ···················· | ···················· |
| P | 8350 | hCoV-19/Denmark/DCGC-6618/2020\|EPI_ISL_619667\|2020 | ···················· | ···················· | ····················· | ···················· | ···················· | ···················· |
| P | 8351 | hCoV-19/Denmark/DCGC-6619/2020\|EPI_ISL_619668\|2020 | ···················· | ···················· | ····················· | ···················· | ···················· | ···················· |
| P | 8352 | hCoV-19/Denmark/DCGC-6621/2020\|EPI_ISL_619670\|2020 | ···················· | ···················· | ····················· | ···················· | ···················· | ···················· |
| P | 8353 | hCoV-19/Denmark/DCGC-6624/2020\|EPI_ISL_619673\|2020 | ···················· | ···················· | ····················· | ···················· | ···················· | ···················· |
| P | 8354 | hCoV-19/Denmark/DCGC-6625/2020\|EPI_ISL_619674\|2020 | ···················· | ···················· | ····················· | ···················· | ···················· | ···················· |
| P | 8355 | hCoV-19/Denmark/DCGC-6626/2020\|EPI_ISL_619675\|2020 | ···················· | ···················· | ····················· | ···················· | ···················· | ···················· |
| P | 8356 | hCoV-19/Denmark/DCGC-6627/2020\|EPI_ISL_619676\|2020 | ···················· | ···················· | ····················· | ···················· | ···················· | ···················· |
| P | 8357 | hCoV-19/Denmark/DCGC-6628/2020\|EPI_ISL_619677\|2020 | ···················· | ···················· | ····················· | ···················· | ···················· | ···················· |
| P | 8358 | hCoV-19/Denmark/DCGC-6629/2020\|EPI_ISL_619678\|2020 | ···················· | ···················· | ····················· | ···················· | ···················· | ···················· |
| P | 8359 | hCoV-19/Denmark/DCGC-6834/2020\|EPI_ISL_619679\|2020 | ···················· | ···················· | ····················· | ···················· | ···················· | ···················· |
| P | 8360 | hCoV-19/Denmark/DCGC-6835/2020\|EPI_ISL_619680\|2020 | ···················· | ···················· | ····················· | ···················· | ···················· | ···················· |
| P | 8361 | hCoV-19/Denmark/DCGC-6837/2020\|EPI_ISL_619682\|2020 | ···················· | ···················· | ····················· | ···················· | ···················· | ···················· |
| P | 8362 | hCoV-19/Denmark/DCGC-6839/2020\|EPI_ISL_619684\|2020 | ···················· | ···················· | ····················· | ···················· | ···················· | ···················· |
| P | 8363 | hCoV-19/Denmark/DCGC-6840/2020\|EPI_ISL_619685\|2020 | ···················· | ···················· | ····················· | ···················· | ···················· | ···················· |
| P | 8364 | hCoV-19/Denmark/DCGC-6841/2020\|EPI_ISL_619686\|2020 | ···················· | ···················· | ····················· | ···················· | ···················· | ···················· |
| P | 8365 | hCoV-19/Denmark/DCGC-6844/2020\|EPI_ISL_619689\|2020 | ···················· | ···················· | ····················· | ···················· | ···················· | ···················· |
| P | 8366 | hCoV-19/Denmark/DCGC-6845/2020\|EPI_ISL_619690\|2020 | ···················· | ···················· | ····················· | ···················· | ···················· | ···················· |
| P | 8367 | hCoV-19/Denmark/DCGC-6846/2020\|EPI_ISL_619691\|2020 | ···················· | ···················· | ····················· | ···················· | ···················· | ···················· |
| P | 8368 | hCoV-19/Denmark/DCGC-6847/2020\|EPI_ISL_619692\|2020 | ···················· | ···················· | ····················· | ···················· | ···················· | ···················· |
| P | 8369 | hCoV-19/Denmark/DCGC-6848/2020\|EPI_ISL_619693\|2020 | ···················· | ···················· | ····················· | ···················· | ···················· | ···················· |
| P | 8370 | hCoV-19/Denmark/DCGC-6849/2020\|EPI_ISL_619694\|2020 | ···················· | ···················· | ····················· | ···················· | ···················· | ···················· |
| P | 8371 | hCoV-19/Denmark/DCGC-6850/2020\|EPI_ISL_619695\|2020 | ···················· | ···················· | ····················· | ···················· | ···················· | ···················· |
| P | 8372 | hCoV-19/Denmark/DCGC-6851/2020\|EPI_ISL_619696\|2020 | ···················· | ···················· | ····················· | ···················· | ···················· | ···················· |
| P | 8373 | hCoV-19/Denmark/DCGC-6852/2020\|EPI_ISL_619697\|2020 | ···················· | ···················· | ····················· | ···················· | ···················· | ···················· |
| P | 8374 | hCoV-19/Denmark/DCGC-6855/2020\|EPI_ISL_619700\|2020 | ···················· | ···················· | ····················· | ···················· | ···················· | ···················· |
| P | 8375 | hCoV-19/Denmark/DCGC-6856/2020\|EPI_ISL_619701\|2020 | ···················· | ···················· | ····················· | ···················· | ···················· | ···················· |
| P | 8376 | hCoV-19/Denmark/DCGC-6857/2020\|EPI_ISL_619702\|2020 | ···················· | ···················· | ····················· | ···················· | ···················· | ···················· |
| P | 8377 | hCoV-19/Denmark/DCGC-6859/2020\|EPI_ISL_619704\|2020 | ···················· | ···················· | ····················· | ···················· | ···················· | ···················· |
| P | 8378 | hCoV-19/Denmark/DCGC-6861/2020\|EPI_ISL_619706\|2020 | ···················· | ···················· | ····················· | ···················· | ···················· | ···················· |
| P | 8379 | hCoV-19/Denmark/DCGC-6862/2020\|EPI_ISL_619707\|2020 | ···················· | ···················· | ····················· | ···················· | ···················· | ···················· |
| P | 8380 | hCoV-19/Denmark/DCGC-6864/2020\|EPI_ISL_619709\|2020 | ···················· | ···················· | ····················· | ···················· | ···················· | ···················· |
| P | 8381 | hCoV-19/Denmark/DCGC-6867/2020\|EPI_ISL_619712\|2020 | ···················· | ···················· | ····················· | ···················· | ···················· | ···················· |
| P | 8382 | hCoV-19/Denmark/DCGC-6868/2020\|EPI_ISL_619713\|2020 | ···················· | ···················· | ····················· | ···················· | ···················· | ···················· |
| P | 8383 | hCoV-19/Denmark/DCGC-6869/2020\|EPI_ISL_619714\|2020 | ···················· | ···················· | ····················· | ···················· | ···················· | ···················· |
| P | 8384 | hCoV-19/Denmark/DCGC-6870/2020\|EPI_ISL_619715\|2020 | ···················· | ···················· | ····················· | ···················· | ···················· | ···················· |
| P | 8385 | hCoV-19/Denmark/DCGC-6871/2020\|EPI_ISL_619716\|2020 | ···················· | ···················· | ····················· | ···················· | ···················· | ···················· |
| P | 8386 | hCoV-19/Denmark/DCGC-6872/2020\|EPI_ISL_619717\|2020 | ···················· | ···················· | ····················· | ···················· | ···················· | ···················· |
| P | 8387 | hCoV-19/Denmark/DCGC-6873/2020\|EPI_ISL_619718\|2020 | ···················· | ···················· | ····················· | ···················· | ···················· | ···················· |
| P | 8388 | hCoV-19/Denmark/DCGC-6874/2020\|EPI_ISL_619719\|2020 | ···················· | ···················· | ····················· | ···················· | ···················· | ···················· |
| P | 8389 | hCoV-19/Denmark/DCGC-6875/2020\|EPI_ISL_619720\|2020 | ···················· | ···················· | ····················· | ···················· | ···················· | ···················· |
| P | 8390 | hCoV-19/Denmark/DCGC-6876/2020\|EPI_ISL_619721\|2020 | ···················· | ·A·················· | ····················· | ···················· | ···················· | ···················· |
| P | 8391 | hCoV-19/Denmark/DCGC-6877/2020\|EPI_ISL_619722\|2020 | ···················· | ···················· | ····················· | ···················· | ···················· | ···················· |
| P | 8392 | hCoV-19/Denmark/DCGC-6878/2020\|EPI_ISL_619723\|2020 | ···················· | ···················· | ····················· | ···················· | ···················· | ···················· |
| P | 8393 | hCoV-19/Denmark/DCGC-6881/2020\|EPI_ISL_619726\|2020 | ···················· | ···················· | ····················· | ···················· | ···················· | ···················· |
| P | 8394 | hCoV-19/Denmark/DCGC-6882/2020\|EPI_ISL_619727\|2020 | ···················· | ···················· | ····················· | ···················· | ···················· | ···················· |
| P | 8395 | hCoV-19/Denmark/DCGC-6883/2020\|EPI_ISL_619728\|2020 | ···················· | ···················· | ····················· | ···················· | ···················· | ···················· |
| P | 8396 | hCoV-19/Denmark/DCGC-6884/2020\|EPI_ISL_619729\|2020 | ···················· | ···················· | ····················· | ···················· | ···················· | ···················· |
| P | 8397 | hCoV-19/Denmark/DCGC-4144/2020\|EPI_ISL_621964\|2020 | ···················· | ···················· | ····················· | ···················· | ···················· | ···················· |
| P | 8398 | hCoV-19/Denmark/DCGC-4145/2020\|EPI_ISL_621965\|2020 | ···················· | ···················· | ····················· | ···················· | ···················· | ···················· |
| P | 8399 | hCoV-19/Denmark/DCGC-4146/2020\|EPI_ISL_621966\|2020 | ···················· | ···················· | ····················· | ···················· | ···················· | ···················· |
| P | 8400 | hCoV-19/Denmark/DCGC-4147/2020\|EPI_ISL_621967\|2020 | ···················· | ···················· | ····················· | ···················· | ···················· | ···················· |
| P | 8401 | hCoV-19/Denmark/DCGC-4149/2020\|EPI_ISL_621969\|2020 | ···················· | ···················· | ····················· | ···················· | ···················· | ···················· |
| P | 8402 | hCoV-19/Denmark/DCGC-4150/2020\|EPI_ISL_621970\|2020 | ···················· | ···················· | ····················· | ···················· | ···················· | ···················· |
| P | 8403 | hCoV-19/Denmark/DCGC-4151/2020\|EPI_ISL_621971\|2020 | ···················· | ···················· | ····················· | ···················· | ···················· | ···················· |
| P | 8404 | hCoV-19/Denmark/DCGC-4152/2020\|EPI_ISL_621972\|2020 | ···················· | ···················· | ····················· | ···················· | ···················· | ···················· |
| P | 8405 | hCoV-19/Denmark/DCGC-4154/2020\|EPI_ISL_621974\|2020 | ···················· | ···················· | ····················· | ···················· | ···················· | ···················· |
| P | 8406 | hCoV-19/Denmark/DCGC-4155/2020\|EPI_ISL_621975\|2020 | ···················· | ···················· | ····················· | ···················· | ···················· | ···················· |
| P | 8407 | hCoV-19/Denmark/DCGC-4156/2020\|EPI_ISL_621976\|2020 | ···················· | ···················· | ····················· | ···················· | ···················· | ···················· |
| P | 8408 | hCoV-19/Denmark/DCGC-4157/2020\|EPI_ISL_621977\|2020 | ···················· | ···················· | ····················· | ···················· | ···················· | ···················· |
| P | 8409 | hCoV-19/Denmark/DCGC-4158/2020\|EPI_ISL_621978\|2020 | ···················· | ···················· | ····················· | ···················· | ···················· | ···················· |
| P | 8410 | hCoV-19/Denmark/DCGC-4159/2020\|EPI_ISL_621979\|2020 | ···················· | ···················· | ····················· | ···················· | ···················· | ···················· |
| P | 8411 | hCoV-19/Denmark/DCGC-4160/2020\|EPI_ISL_621980\|2020 | ···················· | ·A·················· | ····················· | ···················· | ···················· | ···················· |
| P | 8412 | hCoV-19/Denmark/DCGC-4162/2020\|EPI_ISL_621982\|2020 | ···················· | ···················· | ····················· | ···················· | ···················· | ···················· |
| P | 8413 | hCoV-19/Denmark/DCGC-4163/2020\|EPI_ISL_621983\|2020 | ···················· | ···················· | ····················· | ···················· | ···················· | ···················· |
| P | 8414 | hCoV-19/Denmark/DCGC-4164/2020\|EPI_ISL_621984\|2020 | ···················· | ···················· | ····················· | ···················· | ···················· | ···················· |
| P | 8415 | hCoV-19/Denmark/DCGC-4165/2020\|EPI_ISL_621985\|2020 | ···················· | ···················· | ····················· | ···················· | ···················· | ···················· |
| P | 8416 | hCoV-19/Denmark/DCGC-4166/2020\|EPI_ISL_621986\|2020 | ···················· | ···················· | ····················· | ···················· | ···················· | ···················· |
| P | 8417 | hCoV-19/Denmark/DCGC-4167/2020\|EPI_ISL_621987\|2020 | ···················· | ···················· | ····················· | ···················· | ···················· | ···················· |
| P | 8418 | hCoV-19/Denmark/DCGC-4168/2020\|EPI_ISL_621988\|2020 | ···················· | ···················· | ····················· | ···················· | ···················· | ···················· |
| P | 8419 | hCoV-19/Denmark/DCGC-4169/2020\|EPI_ISL_621989\|2020 | ···················· | ···················· | ····················· | ···················· | ···················· | ···················· |
| P | 8420 | hCoV-19/Denmark/DCGC-4170/2020\|EPI_ISL_621990\|2020 | ···················· | ···················· | ····················· | ···················· | ···················· | ···················· |
| P | 8421 | hCoV-19/Denmark/DCGC-4172/2020\|EPI_ISL_621992\|2020 | ···················· | ···················· | ····················· | ···················· | ···················· | ···················· |
| P | 8422 | hCoV-19/Denmark/DCGC-4173/2020\|EPI_ISL_621993\|2020 | ···················· | ···················· | ····················· | ···················· | ···················· | ···················· |
| P | 8423 | hCoV-19/Denmark/DCGC-4175/2020\|EPI_ISL_621995\|2020 | ···················· | ···················· | ····················· | ···················· | ···················· | ···················· |
| P | 8424 | hCoV-19/Denmark/DCGC-4176/2020\|EPI_ISL_621996\|2020 | ···················· | ···················· | ····················· | ···················· | ···················· | ···················· |

P 8425 hCoV-19/Denmark/DCGC-4177/2020|EPI_ISL_621997|2020 ···················· ···················· ···················· ···················· ···················· ····················
P 8426 hCoV-19/Denmark/DCGC-4178/2020|EPI_ISL_621998|2020 ···················· ···················· ···················· ···················· ···················· ····················
P 8427 hCoV-19/Denmark/DCGC-4179/2020|EPI_ISL_621999|2020 ···················· ···················· ···················· ···················· ···················· ····················
P 8428 hCoV-19/England/ALDP-AA48FD/2020|EPI_ISL_624280|20 ···················· ···················· ···················· ···················· ···················· ····················
P 8429 hCoV-19/England/ALDP-AA3BB9/2020|EPI_ISL_624281|20 ···················· ···················· ···················· ···················· ···················· ····················
P 8430 hCoV-19/England/ALDP-AA1CF4/2020|EPI_ISL_624283|20 ···················· ···················· ···················· ···················· ···················· ····················
P 8431 hCoV-19/Wales/ALDP-AA48DF/2020|EPI_ISL_624284|2020 ···················· ···················· ···················· ···················· ···················· ····················
P 8432 hCoV-19/England/ALDP-AA1D1F/2020|EPI_ISL_624285|20 ···················· ···················· ···················· ···················· ···················· ····················
P 8433 hCoV-19/England/ALDP-AA4295/2020|EPI_ISL_624286|20 ···················· ···················· ···················· ···················· ···················· ····················
P 8434 hCoV-19/England/ALDP-AA4259/2020|EPI_ISL_624287|20 ···················· ···················· ···················· ···················· ···················· ····················
P 8435 hCoV-19/England/ALDP-AA3AAD/2020|EPI_ISL_624288|20 ···················· ···················· ···················· ···················· ···················· ····················
P 8436 hCoV-19/England/ALDP-AA423B/2020|EPI_ISL_624289|20 ···················· ···················· ···················· ···················· ···················· ····················
P 8437 hCoV-19/England/ALDP-AA3CF2/2020|EPI_ISL_624290|20 ···················· ···················· ···················· ···················· ···················· ····················
P 8438 hCoV-19/England/ALDP-AA4B7C/2020|EPI_ISL_624292|20 ···················· ···················· ···················· ···················· ···················· ····················
P 8439 hCoV-19/England/ALDP-AA48C0/2020|EPI_ISL_624293|20 ···················· ···················· ···················· ···················· ···················· ····················
P 8440 hCoV-19/England/ALDP-AA42A4/2020|EPI_ISL_624294|20 ···················· ···················· ···················· ···················· ···················· ····················
P 8441 hCoV-19/England/ALDP-AA4C3D/2020|EPI_ISL_624295|20 ···················· ···················· ···················· ···················· ···················· ···A················
P 8442 hCoV-19/England/ALDP-AA422C/2020|EPI_ISL_624296|20 ···················· ···················· ···················· ···················· ···················· ····················
P 8443 hCoV-19/England/ALDP-AA2376/2020|EPI_ISL_624297|20 ···················· ·A·················· ···················· ···················· ···················· ···A················
P 8444 hCoV-19/England/ALDP-AA1AFA/2020|EPI_ISL_624298|20 ···················· ···················· ···················· ···················· ···················· ····················
P 8446 hCoV-19/USA/CA-CZB-11105/2020|EPI_ISL_625501|2020- ···················· ···················· ···················· ···················· ···················· ····················
P 8447 hCoV-19/USA/CA-CZB-11108/2020|EPI_ISL_625502|2020- ···················· ···················· ···················· ···················· ···················· ····················
P 8448 hCoV-19/USA/CA-CZB-11109/2020|EPI_ISL_625503|2020- ···················· ···················· ···················· ···················· ···················· ····················
P 8449 hCoV-19/Denmark/DCGC-1243/2020|EPI_ISL_614759|2020 ···················· ···················· ···················· ···················· ···················· ····················
P 8450 hCoV-19/Denmark/DCGC-1244/2020|EPI_ISL_614760|2020 ···················· ···················· ···················· ···················· ···················· ····················
P 8451 hCoV-19/Denmark/DCGC-759/2020|EPI_ISL_614761|2020- ···················· ···················· ···················· ···················· ···················· ····················
P 8452 hCoV-19/Denmark/DCGC-1245/2020|EPI_ISL_614762|2020 ···················· ···················· ···················· ···················· ···················· ····················
P 8453 hCoV-19/Denmark/DCGC-771/2020|EPI_ISL_614764|2020- ···················· ···················· ···················· ···················· ···················· ····················
P 8454 hCoV-19/Denmark/DCGC-1246/2020|EPI_ISL_614765|2020 ···················· ···················· ···················· ···················· ···················· ····················
P 8455 hCoV-19/Denmark/DCGC-1247/2020|EPI_ISL_614766|2020 ···················· ···················· ···················· ···················· ···················· ····················
P 8456 hCoV-19/Denmark/DCGC-1248/2020|EPI_ISL_614767|2020 ···················· ···················· ···················· ···················· ···················· ····················
P 8457 hCoV-19/Denmark/DCGC-772/2020|EPI_ISL_614768|2020- ···················· ···················· ···················· ···················· ···················· ····················
P 8458 hCoV-19/Denmark/DCGC-1235/2020|EPI_ISL_614769|2020 ···················· ···················· ···················· ···················· ···················· ····················
P 8459 hCoV-19/Denmark/DCGC-760/2020|EPI_ISL_614770|2020- ···················· ···················· ···················· ···················· ···················· ····················
P 8460 hCoV-19/Denmark/DCGC-1236/2020|EPI_ISL_614771|2020 ···················· ···················· ···················· ···················· ···················· ····················
P 8461 hCoV-19/Denmark/DCGC-762/2020|EPI_ISL_614772|2020- ···················· ···················· ···················· ···················· ···················· ····················
P 8462 hCoV-19/Denmark/DCGC-1238/2020|EPI_ISL_614773|2020 ···················· ···················· ···················· ···················· ···················· ····················
P 8463 hCoV-19/Denmark/DCGC-763/2020|EPI_ISL_614774|2020- ···················· ···················· ···················· ···················· ···················· ····················
P 8464 hCoV-19/Denmark/DCGC-766/2020|EPI_ISL_614775|2020- ···················· ···················· ···················· ···················· ···················· ····················
P 8465 hCoV-19/Denmark/DCGC-1003/2020|EPI_ISL_614776|2020 ···················· ···················· ···················· ···················· ···················· ····················
P 8466 hCoV-19/Denmark/DCGC-1006/2020|EPI_ISL_614777|2020 ···················· ···················· ···················· ···················· ···················· ····················
P 8467 hCoV-19/Denmark/DCGC-1007/2020|EPI_ISL_614778|2020 ···················· ···················· ···················· ···················· ···················· ····················
P 8468 hCoV-19/Denmark/DCGC-769/2020|EPI_ISL_614779|2020- ···················· ···················· ···················· ···················· ···················· ····················
P 8469 hCoV-19/Denmark/DCGC-1009/2020|EPI_ISL_614780|2020 ···················· ···················· ···················· ···················· ···················· ····················
P 8470 hCoV-19/Denmark/DCGC-1010/2020|EPI_ISL_614781|2020 ···················· ···················· ···················· ···················· ···················· ····················
P 8471 hCoV-19/Denmark/DCGC-1011/2020|EPI_ISL_614782|2020 ···················· ···················· ···················· ···················· ···················· ····················
P 8472 hCoV-19/Denmark/DCGC-1038/2020|EPI_ISL_614783|2020 ···················· ···················· ···················· ···················· ···················· ····················
P 8473 hCoV-19/Denmark/DCGC-1242/2020|EPI_ISL_614784|2020 ···················· ···················· ···················· ···················· ···················· ····················
P 8474 hCoV-19/Denmark/DCGC-852/2020|EPI_ISL_614785|2020- ···················· ···················· ···················· ···················· ···················· ····················
P 8475 hCoV-19/Denmark/DCGC-870/2020|EPI_ISL_614786|2020- ···················· ···················· ···················· ···················· ···················· ····················
P 8476 hCoV-19/Denmark/DCGC-873/2020|EPI_ISL_614787|2020- ···················· ···················· ···················· ···················· ···················· ····················
P 8477 hCoV-19/Denmark/DCGC-877/2020|EPI_ISL_614788|2020- ···················· ···················· ···················· ···················· ···················· ····················
P 8478 hCoV-19/Denmark/DCGC-849/2020|EPI_ISL_614789|2020- ···················· ···················· ···················· ···················· ···················· ····················
P 8479 hCoV-19/Denmark/DCGC-764/2020|EPI_ISL_614790|2020- ···················· ···················· ···················· ···················· ···················· ····················
P 8480 hCoV-19/Denmark/DCGC-853/2020|EPI_ISL_614791|2020- ···················· ···················· ···················· ···················· ···················· ····················
P 8481 hCoV-19/Denmark/DCGC-863/2020|EPI_ISL_614792|2020- ···················· ···················· ···················· ···················· ···················· ····················
P 8482 hCoV-19/Denmark/DCGC-871/2020|EPI_ISL_614793|2020- ···················· ···················· ···················· ···················· ···················· ····················
P 8483 hCoV-19/Denmark/DCGC-874/2020|EPI_ISL_614794|2020- ···················· ···················· ···················· ···················· ···················· ····················
P 8484 hCoV-19/Denmark/DCGC-854/2020|EPI_ISL_614795|2020- ···················· ···················· ···················· ···················· ···················· ····················
P 8485 hCoV-19/Denmark/DCGC-1239/2020|EPI_ISL_614796|2020 ···················· ···················· ···················· ···················· ···················· ····················
P 8486 hCoV-19/Denmark/DCGC-1241/2020|EPI_ISL_614797|2020 ···················· ···················· ···················· ···················· ···················· ····················
P 8487 hCoV-19/Denmark/DCGC-879/2020|EPI_ISL_614798|2020- ···················· ···················· ···················· ···················· ···················· ····················
P 8488 hCoV-19/Denmark/DCGC-880/2020|EPI_ISL_614799|2020- ···················· ···················· ···················· ···················· ···················· ····················
P 8489 hCoV-19/Denmark/DCGC-884/2020|EPI_ISL_614800|2020- ···················· ···················· ···················· ···················· ···················· ····················
P 8490 hCoV-19/Denmark/DCGC-885/2020|EPI_ISL_614801|2020- ···················· ···················· ···················· ···················· ···················· ····················
P 8491 hCoV-19/Denmark/DCGC-997/2020|EPI_ISL_614802|2020- ···················· ···················· ···················· ···················· ···················· ····················
P 8492 hCoV-19/Denmark/DCGC-998/2020|EPI_ISL_614803|2020- ···················· ···················· ···················· ···················· ···················· ····················
P 8493 hCoV-19/Denmark/DCGC-999/2020|EPI_ISL_614804|2020- ···················· ···················· ···················· ···················· ···················· ····················
P 8494 hCoV-19/Denmark/DCGC-1000/2020|EPI_ISL_614805|2020 ···················· ···················· ···················· ···················· ···················· ····················
P 8495 hCoV-19/Denmark/DCGC-1001/2020|EPI_ISL_614806|2020 ···················· ···················· ···················· ···················· ···················· ····················
P 8496 hCoV-19/Denmark/DCGC-1002/2020|EPI_ISL_614807|2020 ···················· ···················· ···················· ···················· ···················· ····················
P 8497 hCoV-19/Denmark/DCGC-867/2020|EPI_ISL_614808|2020- ···················· ···················· ···················· ···················· ···················· ····················
P 8498 hCoV-19/Denmark/DCGC-869/2020|EPI_ISL_614809|2020- ···················· ···················· ···················· ···················· ···················· ····················
P 8499 hCoV-19/Denmark/DCGC-916/2020|EPI_ISL_614810|2020- ···················· ···················· ···················· ···················· ···················· ····················
P 8500 hCoV-19/Denmark/DCGC-858/2020|EPI_ISL_614811|2020- ···················· ···················· ···················· ···················· ···················· ····················
P 8501 hCoV-19/Denmark/DCGC-861/2020|EPI_ISL_614812|2020- ···················· ···················· ···················· ···················· ···················· ····················
P 8502 hCoV-19/Denmark/DCGC-868/2020|EPI_ISL_614813|2020- ···················· ···················· ···················· ···················· ···················· ····················
P 8503 hCoV-19/Denmark/DCGC-855/2020|EPI_ISL_614814|2020- ···················· ···················· ···················· ···················· ···················· ····················
P 8504 hCoV-19/Denmark/DCGC-850/2020|EPI_ISL_614815|2020- ···················· ···················· ···················· ···················· ···················· ····················
P 8505 hCoV-19/Denmark/DCGC-859/2020|EPI_ISL_614816|2020- ···················· ···················· ···················· ···················· ···················· ····················

| | | | | | | | | |
|---|---|---|---|---|---|---|---|---|
| P | 8506 | hCoV-19/Denmark/DCGC-864/2020\|EPI_ISL_614817\|2020- | ···················· | ···················· | ····················· | ···················· | ···················· | ···················· |
| P | 8507 | hCoV-19/Denmark/DCGC-878/2020\|EPI_ISL_614818\|2020- | ···················· | ···················· | ····················· | ···················· | ···················· | ···················· |
| P | 8508 | hCoV-19/Denmark/DCGC-851/2020\|EPI_ISL_614819\|2020- | ···················· | ···················· | ····················· | ···················· | ···················· | ···················· |
| P | 8509 | hCoV-19/Denmark/DCGC-856/2020\|EPI_ISL_614820\|2020- | ···················· | ···················· | ····················· | ···················· | ···················· | ···················· |
| P | 8510 | hCoV-19/Denmark/DCGC-915/2020\|EPI_ISL_614821\|2020- | ···················· | ···················· | ····················· | ···················· | ···················· | ···················· |
| P | 8511 | hCoV-19/Denmark/DCGC-865/2020\|EPI_ISL_614822\|2020- | ···················· | ···················· | ····················· | ···················· | ···················· | ···················· |
| P | 8512 | hCoV-19/Denmark/DCGC-917/2020\|EPI_ISL_614823\|2020- | ···················· | ···················· | ····················· | ···················· | ···················· | ···················· |
| P | 8513 | hCoV-19/Denmark/DCGC-876/2020\|EPI_ISL_614824\|2020- | ···················· | ···················· | ····················· | ···················· | ···················· | ···················· |
| P | 8514 | hCoV-19/Denmark/DCGC-918/2020\|EPI_ISL_614825\|2020- | ···················· | ···················· | ····················· | ···················· | ···················· | ···················· |
| P | 8515 | hCoV-19/Denmark/DCGC-857/2020\|EPI_ISL_614826\|2020- | ···················· | ···················· | ····················· | ···················· | ···················· | ···················· |
| P | 8516 | hCoV-19/Denmark/DCGC-860/2020\|EPI_ISL_614827\|2020- | ···················· | ···················· | ····················· | ···················· | ···················· | ···················· |
| P | 8517 | hCoV-19/Denmark/DCGC-866/2020\|EPI_ISL_614828\|2020- | ···················· | ···················· | ····················· | ···················· | ···················· | ···················· |
| P | 8518 | hCoV-19/Denmark/DCGC-872/2020\|EPI_ISL_614829\|2020- | ···················· | ···················· | ····················· | ···················· | ···················· | ···················· |
| P | 8519 | hCoV-19/Denmark/DCGC-1184/2020\|EPI_ISL_614830\|2020 | ···················· | ···················· | ····················· | ···················· | ···················· | ···················· |
| P | 8520 | hCoV-19/Denmark/DCGC-1185/2020\|EPI_ISL_614831\|2020 | ···················· | ···················· | ····················· | ···················· | ···················· | ···················· |
| P | 8521 | hCoV-19/Denmark/DCGC-1186/2020\|EPI_ISL_614832\|2020 | ···················· | ···················· | ····················· | ···················· | ···················· | ···················· |
| P | 8522 | hCoV-19/Denmark/DCGC-1187/2020\|EPI_ISL_614833\|2020 | ···················· | ···················· | ····················· | ···················· | ···················· | ···················· |
| P | 8523 | hCoV-19/Denmark/DCGC-1188/2020\|EPI_ISL_614834\|2020 | ···················· | ···················· | ····················· | ···················· | ···················· | ···················· |
| P | 8524 | hCoV-19/Denmark/DCGC-1189/2020\|EPI_ISL_614835\|2020 | ···················· | ···················· | ····················· | ···················· | ···················· | ···················· |
| P | 8525 | hCoV-19/Denmark/DCGC-1202/2020\|EPI_ISL_614847\|2020 | ···················· | ···················· | ····················· | ···················· | ···················· | ···················· |
| P | 8526 | hCoV-19/Denmark/DCGC-1203/2020\|EPI_ISL_614848\|2020 | ···················· | ···················· | ····················· | ···················· | ···················· | ···················· |
| P | 8527 | hCoV-19/Denmark/DCGC-1204/2020\|EPI_ISL_614849\|2020 | ···················· | ···················· | ····················· | ···················· | ···················· | ···················· |
| P | 8528 | hCoV-19/Denmark/DCGC-1209/2020\|EPI_ISL_614854\|2020 | ···················· | ···················· | ····················· | ···················· | ···················· | ···················· |
| P | 8529 | hCoV-19/Denmark/DCGC-1211/2020\|EPI_ISL_614855\|2020 | ···················· | ···················· | ····················· | ···················· | ···················· | ···················· |
| P | 8530 | hCoV-19/Denmark/DCGC-1215/2020\|EPI_ISL_614858\|2020 | ···················· | ···················· | ····················· | ···················· | ···················· | ···················· |
| P | 8531 | hCoV-19/Denmark/DCGC-1216/2020\|EPI_ISL_614859\|2020 | ···················· | ···················· | ····················· | ···················· | ···················· | ···················· |
| P | 8532 | hCoV-19/Denmark/DCGC-1217/2020\|EPI_ISL_614860\|2020 | ···················· | ···················· | ····················· | ···················· | ···················· | ···················· |
| P | 8533 | hCoV-19/Denmark/DCGC-1218/2020\|EPI_ISL_614861\|2020 | ···················· | ···················· | ····················· | ···················· | ···················· | ···················· |
| P | 8534 | hCoV-19/Denmark/DCGC-1219/2020\|EPI_ISL_614862\|2020 | ···················· | ···················· | ····················· | ···················· | ···················· | ···················· |
| P | 8535 | hCoV-19/Denmark/DCGC-1220/2020\|EPI_ISL_614863\|2020 | ···················· | ···················· | ····················· | ···················· | ···················· | ···················· |
| P | 8536 | hCoV-19/Denmark/DCGC-1221/2020\|EPI_ISL_614864\|2020 | ···················· | ···················· | ····················· | ···················· | ···················· | ···················· |
| P | 8537 | hCoV-19/Denmark/DCGC-1222/2020\|EPI_ISL_614865\|2020 | ···················· | ···················· | ····················· | ···················· | ···················· | ···················· |
| P | 8538 | hCoV-19/Denmark/DCGC-1223/2020\|EPI_ISL_614866\|2020 | ···················· | ···················· | ····················· | ···················· | ···················· | ···················· |
| P | 8539 | hCoV-19/Denmark/DCGC-1224/2020\|EPI_ISL_614867\|2020 | ···················· | ···················· | ····················· | ···················· | ···················· | ···················· |
| P | 8540 | hCoV-19/Denmark/DCGC-1225/2020\|EPI_ISL_614868\|2020 | ···················· | ···················· | ····················· | ···················· | ···················· | ···················· |
| P | 8541 | hCoV-19/Denmark/DCGC-1226/2020\|EPI_ISL_614869\|2020 | ···················· | ···················· | ····················· | ···················· | ···················· | ···················· |
| P | 8542 | hCoV-19/Denmark/DCGC-1229/2020\|EPI_ISL_614870\|2020 | ···················· | ···················· | ····················· | ···················· | ···················· | ···················· |
| P | 8543 | hCoV-19/Denmark/DCGC-1230/2020\|EPI_ISL_614871\|2020 | ···················· | ···················· | ····················· | ···················· | ···················· | ···················· |
| P | 8544 | hCoV-19/Denmark/DCGC-1231/2020\|EPI_ISL_614872\|2020 | ···················· | ···················· | ····················· | ···················· | ···················· | ···················· |
| P | 8545 | hCoV-19/Denmark/DCGC-1232/2020\|EPI_ISL_614873\|2020 | ···················· | ···················· | ····················· | ···················· | ···················· | ···················· |
| P | 8546 | hCoV-19/Denmark/DCGC-1233/2020\|EPI_ISL_614874\|2020 | ···················· | ···················· | ····················· | ···················· | ···················· | ···················· |
| P | 8547 | hCoV-19/Denmark/DCGC-1234/2020\|EPI_ISL_614875\|2020 | ···················· | ···················· | ····················· | ···················· | ···················· | ···················· |
| P | 8548 | hCoV-19/Denmark/DCGC-752/2020\|EPI_ISL_614876\|2020- | ···················· | ···················· | ····················· | ···················· | ···················· | ···················· |
| P | 8549 | hCoV-19/Denmark/DCGC-754/2020\|EPI_ISL_614877\|2020- | ···················· | ···················· | ····················· | ···················· | ···················· | ···················· |
| P | 8550 | hCoV-19/Denmark/DCGC-775/2020\|EPI_ISL_614878\|2020- | ···················· | ···················· | ····················· | ···················· | ···················· | ···················· |
| P | 8551 | hCoV-19/Denmark/DCGC-756/2020\|EPI_ISL_614879\|2020- | ···················· | ···················· | ····················· | ···················· | ···················· | ···················· |
| P | 8552 | hCoV-19/Denmark/DCGC-8028/2020\|EPI_ISL_617077\|2020 | ···················· | ···················· | ····················· | ···················· | ···················· | ···················· |
| P | 8553 | hCoV-19/Denmark/DCGC-8123/2020\|EPI_ISL_617078\|2020 | ···················· | ···················· | ····················· | ···················· | ···················· | ···················· |
| P | 8554 | hCoV-19/Denmark/DCGC-7923/2020\|EPI_ISL_617096\|2020 | ···················· | ···················· | ····················· | ···················· | ···················· | ···················· |
| P | 8555 | hCoV-19/Denmark/DCGC-7930/2020\|EPI_ISL_617097\|2020 | ···················· | ···················· | ····················· | ···················· | ···················· | ···················· |
| P | 8556 | hCoV-19/Denmark/DCGC-7961/2020\|EPI_ISL_617098\|2020 | ···················· | ···················· | ····················· | ···················· | ···················· | ···················· |
| P | 8557 | hCoV-19/Denmark/DCGC-7931/2020\|EPI_ISL_617099\|2020 | ···················· | ···················· | ····················· | ···················· | ···················· | ···················· |
| P | 8558 | hCoV-19/Denmark/DCGC-6565/2020\|EPI_ISL_619613\|2020 | ···················· | ···················· | ····················· | ···················· | ···················· | ···················· |
| P | 8559 | hCoV-19/Denmark/DCGC-6566/2020\|EPI_ISL_619614\|2020 | ···················· | ···················· | ····················· | ···················· | ···················· | ···················· |
| P | 8560 | hCoV-19/Denmark/DCGC-6567/2020\|EPI_ISL_619615\|2020 | ···················· | ···················· | ····················· | ···················· | ···················· | ···················· |
| P | 8561 | hCoV-19/Denmark/DCGC-6568/2020\|EPI_ISL_619616\|2020 | ···················· | ···················· | ····················· | ···················· | ···················· | ···················· |
| P | 8562 | hCoV-19/Denmark/DCGC-6571/2020\|EPI_ISL_619619\|2020 | ···················· | ···················· | ····················· | ···················· | ···················· | ···················· |
| P | 8563 | hCoV-19/Denmark/DCGC-4700/2020\|EPI_ISL_621932\|2020 | ···················· | ···················· | ····················· | ···················· | ···················· | ···················· |
| P | 8564 | hCoV-19/Denmark/DCGC-4702/2020\|EPI_ISL_621933\|2020 | ···················· | ···················· | ····················· | ···················· | ···················· | ···················· |
| P | 8565 | hCoV-19/Denmark/DCGC-4704/2020\|EPI_ISL_621935\|2020 | ···················· | ···················· | ····················· | ···················· | ···················· | ···················· |
| P | 8566 | hCoV-19/Denmark/DCGC-4705/2020\|EPI_ISL_621936\|2020 | ···················· | ···················· | ····················· | ···················· | ···················· | ···················· |
| P | 8567 | hCoV-19/England/ALDP-AA51A6/2020\|EPI_ISL_624100\|20 | ···················· | ···················· | ····················· | ···················· | ···················· | ···················· |
| P | 8568 | hCoV-19/England/ALDP-AA2E57/2020\|EPI_ISL_624102\|20 | ···················· | ···················· | ····················· | ···················· | ···················· | ···················· |
| P | 8569 | hCoV-19/England/ALDP-AA12F2/2020\|EPI_ISL_624103\|20 | ···················· | ···················· | ····················· | ···················· | ···················· | ···················· |
| P | 8570 | hCoV-19/England/ALDP-AA2ABD/2020\|EPI_ISL_624104\|20 | ···················· | ···················· | ····················· | ···················· | ···················· | ···················· |
| P | 8571 | hCoV-19/Scotland/QEUH-AADC34/2020\|EPI_ISL_624105\|2 | ···················· | ···················· | ····················· | ···················· | ····T··············· | ···················· |
| P | 8572 | hCoV-19/England/ALDP-AA1966/2020\|EPI_ISL_624106\|20 | ···················· | ···················· | ····················· | ···················· | ···················· | ···················· |
| P | 8573 | hCoV-19/England/ALDP-AA41E3/2020\|EPI_ISL_624107\|20 | ···················· | ···················· | ····················· | ···················· | ···················· | ···················· |
| P | 8574 | hCoV-19/England/ALDP-AA2EEE/2020\|EPI_ISL_624108\|20 | ···················· | ···················· | ····················· | ···················· | ···················· | ···················· |
| P | 8575 | hCoV-19/England/ALDP-AA2B32/2020\|EPI_ISL_624109\|20 | ···················· | ·A·················· | ····················· | ···················· | ···················· | ··A················· |
| P | 8576 | hCoV-19/England/ALDP-AA0DF2/2020\|EPI_ISL_624110\|20 | ···················· | ···················· | ····················· | ···················· | ···················· | ···················· |
| P | 8577 | hCoV-19/England/ALDP-AA56D4/2020\|EPI_ISL_624111\|20 | ···················· | ···················· | ····················· | ···················· | ···················· | ···················· |
| P | 8578 | hCoV-19/Scotland/QEUH-AAEEF1/2020\|EPI_ISL_624112\|2 | ···················· | ···················· | ····················· | ···················· | ···················· | ···················· |
| P | 8579 | hCoV-19/England/ALDP-AA3533/2020\|EPI_ISL_624113\|20 | ···················· | ···················· | ····················· | ···················· | ···················· | ···················· |
| P | 8580 | hCoV-19/Scotland/QEUH-AAE4C2/2020\|EPI_ISL_624114\|2 | ···················· | ···················· | ····················· | ···················· | ···················· | ···················· |
| P | 8581 | hCoV-19/England/ALDP-AA3111/2020\|EPI_ISL_624115\|20 | ···················· | ···················· | ····················· | ···················· | ···················· | ···················· |
| P | 8582 | hCoV-19/England/ALDP-AA1368/2020\|EPI_ISL_624116\|20 | ···················· | ···················· | ····················· | ···················· | ···················· | ···················· |
| P | 8583 | hCoV-19/England/ALDP-AA18E1/2020\|EPI_ISL_624117\|20 | ···················· | ···················· | ····················· | ···················· | ···················· | ···················· |
| P | 8584 | hCoV-19/England/ALDP-AA358E/2020\|EPI_ISL_624118\|20 | ···················· | ···················· | ····················· | ···················· | ···················· | ···················· |
| P | 8585 | hCoV-19/England/QEUH-AAEF3A/2020\|EPI_ISL_624120\|20 | ···················· | ···················· | ····················· | ···················· | ···················· | ···················· |

| | | | | | | | |
|---|---|---|---|---|---|---|---|
| P | 8586 hCoV-19/England/ALDP-AA35AC/2020\|EPI_ISL_624121\|20 | ···················· | ···················· | ···················· | ···················· | ···················· | ···················· |
| P | 8587 hCoV-19/England/QEUH-AADD13/2020\|EPI_ISL_624122\|20 | ···················· | ···················· | ···················· | ···················· | ···················· | ···················· |
| P | 8588 hCoV-19/England/ALDP-AA331B/2020\|EPI_ISL_624124\|20 | ···················· | ···················· | ···················· | ···················· | ···················· | ···················· |
| P | 8589 hCoV-19/England/ALDP-AA409B/2020\|EPI_ISL_624125\|20 | ···················· | ···················· | ···················· | ···················· | ···················· | ···················· |
| P | 8590 hCoV-19/England/ALDP-AA375B/2020\|EPI_ISL_624126\|20 | ···················· | ···················· | ···················· | ···················· | ···················· | ···················· |
| P | 8591 hCoV-19/England/ALDP-AA124D/2020\|EPI_ISL_624127\|20 | ···················· | ···················· | ···················· | ···················· | ···················· | ···················· |
| P | 8592 hCoV-19/England/ALDP-AA0E86/2020\|EPI_ISL_624129\|20 | ···················· | ·A·················· | ···················· | ···················· | ···················· | ··A················· |
| P | 8593 hCoV-19/England/ALDP-AA2E48/2020\|EPI_ISL_624130\|20 | ···················· | ···················· | ···················· | ···················· | ···················· | ···················· |
| P | 8594 hCoV-19/Scotland/QEUH-AADCF8/2020\|EPI_ISL_624132\|2 | ···················· | ···················· | ···················· | ···················· | ···················· | ···················· |
| P | 8595 hCoV-19/England/ALDP-AA40F5/2020\|EPI_ISL_624134\|20 | ···················· | ···················· | ···················· | ···················· | ···················· | ···················· |
| P | 8596 hCoV-19/England/ALDP-AA3788/2020\|EPI_ISL_624136\|20 | ···················· | ···················· | ···················· | ···················· | ···················· | ···················· |
| P | 8597 hCoV-19/England/ALDP-AA0DA7/2020\|EPI_ISL_624137\|20 | ···················· | ···················· | ···················· | ···················· | ···················· | ···················· |
| P | 8598 hCoV-19/England/ALDP-AA2B41/2020\|EPI_ISL_624138\|20 | ···················· | ···················· | ···················· | ···················· | ···················· | ···················· |
| P | 8599 hCoV-19/England/ALDP-AA2E93/2020\|EPI_ISL_624139\|20 | ···················· | ···················· | ···················· | ···················· | ···················· | ···················· |
| P | 8600 hCoV-19/Scotland/QEUH-AAE50B/2020\|EPI_ISL_624140\|2 | ···················· | ···················· | ···················· | ···················· | ···················· | ···················· |
| P | 8601 hCoV-19/England/ALDP-AA521C/2020\|EPI_ISL_624141\|20 | ···················· | ···················· | ···················· | ···················· | ···················· | ···················· |
| P | 8602 hCoV-19/England/ALDP-AA30F6/2020\|EPI_ISL_624142\|20 | ···················· | ···················· | ···················· | ···················· | ···················· | ···················· |
| P | 8603 hCoV-19/England/ALDP-AA41D4/2020\|EPI_ISL_624143\|20 | ···················· | ···················· | ···················· | ···················· | ···················· | ···················· |
| P | 8604 hCoV-19/England/ALDP-AA2EA2/2020\|EPI_ISL_624144\|20 | ···················· | ···················· | ···················· | ···················· | ···················· | ···················· |
| P | 8605 hCoV-19/Scotland/QEUH-AAE88A/2020\|EPI_ISL_624145\|2 | ···················· | ···················· | ···················· | ···················· | ···················· | ···················· |
| P | 8606 hCoV-19/England/ALDP-AA407D/2020\|EPI_ISL_624146\|20 | ···················· | ···················· | ···················· | ···················· | ···················· | ···················· |
| P | 8607 hCoV-19/Scotland/QEUH-AAD3C6/2020\|EPI_ISL_624147\|2 | ···················· | ···················· | ···················· | ···················· | ···················· | ···················· |
| P | 8608 hCoV-19/Scotland/QEUH-AADC61/2020\|EPI_ISL_624148\|2 | ···················· | ···················· | ···················· | ···················· | ···················· | ···················· |
| P | 8609 hCoV-19/England/ALDP-AA0721/2020\|EPI_ISL_624149\|20 | ···················· | ···················· | ···················· | ···················· | ······T············· | ···················· |
| P | 8610 hCoV-19/England/ALDP-AA3393/2020\|EPI_ISL_624150\|20 | ···················· | ···················· | ···················· | ···················· | ···················· | ···················· |
| P | 8611 hCoV-19/England/ALDP-AA0E4A/2020\|EPI_ISL_624151\|20 | ···················· | ·A·················· | ···················· | ···················· | ···················· | ··A················· |
| P | 8612 hCoV-19/England/ALDP-AA2D69/2020\|EPI_ISL_624152\|20 | ···················· | ···················· | ···················· | ···················· | ···················· | ···················· |
| P | 8613 hCoV-19/England/ALDP-AA417A/2020\|EPI_ISL_624153\|20 | ···················· | ···················· | ···················· | ···················· | ···················· | ···················· |
| P | 8614 hCoV-19/England/ALDP-AA40D7/2020\|EPI_ISL_624154\|20 | ···················· | ···················· | ···················· | ···················· | ···················· | ···················· |
| P | 8615 hCoV-19/England/ALDP-AA314E/2020\|EPI_ISL_624155\|20 | ···················· | ···················· | ···················· | ···················· | ···················· | ···················· |
| P | 8616 hCoV-19/England/ALDP-AA1483/2020\|EPI_ISL_624156\|20 | ···················· | ···················· | ···················· | ···················· | ···················· | ···················· |
| P | 8617 hCoV-19/England/ALDP-AA567A/2020\|EPI_ISL_624157\|20 | ···················· | ···················· | ···················· | ···················· | ···················· | ···················· |
| P | 8618 hCoV-19/England/ALDP-AA3366/2020\|EPI_ISL_624159\|20 | ···················· | ···················· | ···················· | ···················· | ···················· | ···················· |
| P | 8619 hCoV-19/Wales/MILK-A6B106/2020\|EPI_ISL_625407\|2020 | ···················· | ···················· | ···················· | ···················· | ···················· | ···················· |
| P | 8620 hCoV-19/Scotland/MILK-A69DD7/2020\|EPI_ISL_625409\|2 | ···················· | ···················· | ···················· | ···················· | ···················· | ···················· |
| P | 8621 hCoV-19/Scotland/MILK-A69E01/2020\|EPI_ISL_625410\|2 | ···················· | ···················· | ···················· | ···················· | ···················· | ···················· |
| P | 8622 hCoV-19/Scotland/MILK-A69E5C/2020\|EPI_ISL_625411\|2 | ···················· | ···················· | ···················· | ···················· | ···················· | ···················· |
| P | 8623 hCoV-19/Scotland/MILK-A6B77D/2020\|EPI_ISL_625412\|2 | ···················· | ···················· | ···················· | ···················· | ···················· | ···················· |
| P | 8624 hCoV-19/Scotland/MILK-A69E6B/2020\|EPI_ISL_625413\|2 | ···················· | ···················· | ···················· | ···················· | ···················· | ···················· |
| P | 8625 hCoV-19/Wales/MILK-A6B5A0/2020\|EPI_ISL_625414\|2020 | ···················· | ···················· | ···················· | ···················· | ···················· | ···················· |
| P | 8626 hCoV-19/Wales/MILK-A6ABCD/2020\|EPI_ISL_625415\|2020 | ···················· | ···················· | ···················· | ···················· | ···················· | ···················· |
| P | 8627 hCoV-19/Scotland/MILK-A6B063/2020\|EPI_ISL_625416\|2 | ···················· | ···················· | ···················· | ···················· | ···················· | ···················· |
| P | 8628 hCoV-19/Wales/MILK-A6AC60/2020\|EPI_ISL_625417\|2020 | ···················· | ···················· | ···················· | ···················· | ···················· | ···················· |
| P | 8629 hCoV-19/Scotland/MILK-A69E7A/2020\|EPI_ISL_625418\|2 | ···················· | ···················· | ···················· | ···················· | ···················· | ···················· |
| P | 8630 hCoV-19/Scotland/MILK-A69E10/2020\|EPI_ISL_625419\|2 | ···················· | ···················· | ···················· | ···················· | ···················· | ···················· |
| P | 8631 hCoV-19/Scotland/MILK-A6B7B9/2020\|EPI_ISL_625420\|2 | ···················· | ···················· | ···················· | ···················· | ···················· | ···················· |
| P | 8632 hCoV-19/Scotland/MILK-A6B7C8/2020\|EPI_ISL_625421\|2 | ···················· | ···················· | ···················· | ···················· | ···················· | ···················· |
| P | 8633 hCoV-19/Wales/ALDP-A67E6D/2020\|EPI_ISL_625422\|2020 | ···················· | ···················· | ···················· | ···················· | ···················· | ···················· |
| P | 8634 hCoV-19/Scotland/ALDP-A6951B/2020\|EPI_ISL_625423\|2 | ···················· | ···················· | ···················· | ···················· | ···················· | ···················· |
| P | 8635 hCoV-19/Scotland/MILK-A6BDA8/2020\|EPI_ISL_625424\|2 | ···················· | ···················· | ···················· | ···················· | ···················· | ···················· |
| P | 8636 hCoV-19/Wales/ALDP-A67E7C/2020\|EPI_ISL_625425\|2020 | ···················· | ···················· | ···················· | ···················· | ···················· | ···················· |
| P | 8637 hCoV-19/Scotland/MILK-A6BD99/2020\|EPI_ISL_625426\|2 | ···················· | ···················· | ···················· | ···················· | ···················· | ···················· |
| P | 8638 hCoV-19/Wales/ALDP-A67E9A/2020\|EPI_ISL_625427\|2020 | ···················· | ···················· | ···················· | ···················· | ···················· | ···················· |
| P | 8639 hCoV-19/Wales/ALDP-A67EA9/2020\|EPI_ISL_625429\|2020 | ···················· | ···················· | ···················· | ···················· | ···················· | ···················· |
| P | 8640 hCoV-19/Scotland/MILK-A6C0CC/2020\|EPI_ISL_625430\|2 | ···················· | ···················· | ···················· | ···················· | ···················· | ···················· |
| P | 8641 hCoV-19/Wales/ALDP-A67BEE/2020\|EPI_ISL_625431\|2020 | ···················· | ···················· | ···················· | ···················· | ···················· | ···················· |
| P | 8642 hCoV-19/Wales/CAMC-A64E7F/2020\|EPI_ISL_625432\|2020 | ···················· | ···················· | ···················· | ···················· | ···················· | ···················· |
| P | 8643 hCoV-19/Wales/ALDP-A67BB1/2020\|EPI_ISL_625433\|2020 | ···················· | ···················· | ···················· | ···················· | ···················· | ···················· |
| P | 8644 hCoV-19/Wales/ALDP-A67B48/2020\|EPI_ISL_625434\|2020 | ···················· | ···················· | ···················· | ···················· | ···················· | ···················· |
| P | 8645 hCoV-19/Wales/ALDP-A683C7/2020\|EPI_ISL_625435\|2020 | ···················· | ···················· | ···················· | ···················· | ···················· | ···················· |
| P | 8646 hCoV-19/Wales/ALDP-A67BC0/2020\|EPI_ISL_625436\|2020 | ···················· | ···················· | ···················· | ···················· | ···················· | ···················· |
| P | 8647 hCoV-19/Scotland/CAMC-A419C2/2020\|EPI_ISL_625437\|2 | ···················· | ···················· | ···················· | ···················· | ···················· | ···················· |
| P | 8648 hCoV-19/Wales/CAMC-A410DC/2020\|EPI_ISL_625438\|2020 | ···················· | ···················· | ···················· | ···················· | ···················· | ···················· |
| P | 8649 hCoV-19/Wales/CAMC-A418D4/2020\|EPI_ISL_625439\|2020 | ···················· | ···················· | ···················· | ···················· | ···················· | ···················· |
| P | 8650 hCoV-19/Wales/CAMC-A410EB/2020\|EPI_ISL_625440\|2020 | ···················· | ···················· | ···················· | ···················· | ···················· | ···················· |
| P | 8651 hCoV-19/Wales/CAMC-A4179B/2020\|EPI_ISL_625441\|2020 | ···················· | ···················· | ···················· | ···················· | ···················· | ···················· |
| P | 8652 hCoV-19/Wales/CAMC-A4176E/2020\|EPI_ISL_625442\|2020 | ···················· | ···················· | ···················· | ···················· | ···················· | ···················· |
| P | 8653 hCoV-19/Wales/CAMC-A41BEA/2020\|EPI_ISL_625443\|2020 | ···················· | ···················· | ···················· | ···················· | ···················· | ···················· |
| P | 8654 hCoV-19/Wales/CAMC-A3E6C1/2020\|EPI_ISL_625444\|2020 | ···················· | ···················· | ···················· | ···················· | ···················· | ···················· |
| P | 8655 hCoV-19/Scotland/CAMC-A3F736/2020\|EPI_ISL_625446\|2 | ···················· | ···················· | ···················· | ···················· | ···················· | ···················· |
| P | 8656 hCoV-19/Scotland/CAMC-A3F1A1/2020\|EPI_ISL_625447\|2 | ···················· | ···················· | ···················· | ···················· | ···················· | ···················· |
| P | 8657 hCoV-19/Scotland/CAMC-A3F763/2020\|EPI_ISL_625448\|2 | ···················· | ···················· | ···················· | ···················· | ···················· | ···················· |
| P | 8658 hCoV-19/Wales/CAMC-A3E95F/2020\|EPI_ISL_625449\|2020 | ···················· | ···················· | ···················· | ···················· | ···················· | ···················· |
| P | 8659 hCoV-19/Scotland/CAMC-A3F718/2020\|EPI_ISL_625450\|2 | ···················· | ···················· | ···················· | ···················· | ···················· | ···················· |
| P | 8660 hCoV-19/Scotland/CAMC-A3E588/2020\|EPI_ISL_625451\|2 | ···················· | ···················· | ···················· | ···················· | ···················· | ···················· |
| P | 8661 hCoV-19/Scotland/CAMC-A3F0D1/2020\|EPI_ISL_625452\|2 | ···················· | ·A·················· | ···················· | ···················· | ···················· | ···················· |
| P | 8662 hCoV-19/Wales/CAMC-A3E6A3/2020\|EPI_ISL_625453\|2020 | ···················· | ···················· | ···················· | ···················· | ···················· | ···················· |
| P | 8663 hCoV-19/Madagascar/VIRO-5815/2020\|EPI_ISL_625456\|2 | ···················· | ···················· | ···················· | ···················· | ···················· | ···················· |
| P | 8664 hCoV-19/Bangladesh/CHRF-0046/2020\|EPI_ISL_625460\|2 | ···················· | ···················· | ···················· | ···················· | ···················· | ···················· |
| P | 8666 hCoV-19/Bangladesh/CHRF-0051/2020\|EPI_ISL_625465\|2 | ···················· | ···················· | ···················· | ···················· | ···················· | ···················· |

| | | | | | | | | |
|---|---|---|---|---|---|---|---|---|
| P | 8667 hCoV-19/Bangladesh/CHRF-0053/2020\|EPI_ISL_625467\|2 | ···················· | ···················· | ···················· | ···················· | NNN················· | ···················· | 1 |
| P | 8668 hCoV-19/Bangladesh/CHRF-0058/2020\|EPI_ISL_625471\|2 | ···················· | ···················· | ···················· | ···················· | ···················· | ···················· | |
| P | 8669 hCoV-19/Bangladesh/CHRF-0060/2020\|EPI_ISL_625473\|2 | ···················· | ···················· | ···················· | ···················· | ···················· | ···················· | |
| P | 8670 hCoV-19/Bangladesh/CHRF-0061/2020\|EPI_ISL_625474\|2 | ···················· | ···················· | ···················· | ···················· | ···················· | ···················· | |
| P | 8671 hCoV-19/Bangladesh/CHRF-0062/2020\|EPI_ISL_625475\|2 | ···················· | ···················· | ···················· | ···················· | ···················· | ···················· | |
| P | 8673 hCoV-19/Bangladesh/CHRF-0064/2020\|EPI_ISL_625477\|2 | ···················· | ···················· | ···················· | ···················· | ···················· | ···················· | |
| P | 8674 hCoV-19/USA/CA-CZB-11644/2020\|EPI_ISL_625478\|2020- | ···················· | ·A·················· | ···················· | ···················· | ···················· | ···················· | |
| P | 8675 hCoV-19/USA/CA-CZB-11646/2020\|EPI_ISL_625479\|2020- | ···················· | ·A·················· | ···················· | ···················· | ···················· | ···················· | |
| P | 8676 hCoV-19/USA/CA-CZB-11649/2020\|EPI_ISL_625480\|2020- | ···················· | ·A·················· | ···················· | ···················· | ···················· | ···················· | |
| P | 8678 hCoV-19/USA/CA-CZB-11656/2020\|EPI_ISL_625482\|2020- | ···················· | ·A·················· | ···················· | ···················· | ···················· | ···················· | |
| P | 8679 hCoV-19/USA/CA-CZB-11666/2020\|EPI_ISL_625484\|2020- | ···················· | ···················· | ··········T········· | ···················· | ···················· | ···················· | |
| P | 8680 hCoV-19/USA/CA-CZB-11670/2020\|EPI_ISL_625485\|2020- | ···················· | ·A·················· | ···················· | ···················· | ···················· | ···················· | |
| P | 8681 hCoV-19/USA/CA-CZB-11673/2020\|EPI_ISL_625486\|2020- | ···················· | ···················· | ···················· | ···················· | ···················· | ···················· | |
| P | 8682 hCoV-19/USA/CA-CZB-11674/2020\|EPI_ISL_625487\|2020- | ···················· | ·A·················· | ···················· | ···················· | ···················· | ···················· | |
| P | 8683 hCoV-19/USA/CA-CZB-11675/2020\|EPI_ISL_625488\|2020- | ···················· | ···················· | ···················· | ···················· | ···················· | ···················· | |
| P | 8684 hCoV-19/USA/CA-CZB-11677/2020\|EPI_ISL_625489\|2020- | ···················· | ···················· | ···················· | ···················· | ···················· | ···················· | |
| P | 8685 hCoV-19/USA/CA-CZB-11113/2020\|EPI_ISL_625506\|2020- | ···················· | ···················· | ···················· | ···················· | ···················· | ···················· | |
| P | 8686 hCoV-19/USA/CA-CZB-11114/2020\|EPI_ISL_625507\|2020- | ···················· | ···················· | ···················· | ···················· | ···················· | ···················· | |
| P | 8690 hCoV-19/USA/CA-CZB-11118/2020\|EPI_ISL_625511\|2020- | ···················· | ···················· | ···················· | ···················· | ···················· | ···················· | |
| P | 8692 hCoV-19/USA/CA-CZB-11122/2020\|EPI_ISL_625514\|2020- | ···················· | ···················· | ···················· | ···················· | ···················· | ···················· | |
| P | 8694 hCoV-19/USA/CA-CZB-11125/2020\|EPI_ISL_625516\|2020- | ···················· | ···················· | ···················· | ···················· | ···················· | ···················· | |
| P | 8697 hCoV-19/USA/CA-CZB-11129/2020\|EPI_ISL_625519\|2020- | ···················· | ···················· | ···················· | ···················· | ···················· | ···················· | |
| P | 8699 hCoV-19/USA/CA-CZB-11132/2020\|EPI_ISL_625521\|2020- | ···················· | ···················· | ···················· | ···················· | ···················· | ···················· | |
| P | 8700 hCoV-19/USA/CA-CZB-11165/2020\|EPI_ISL_625523\|2020- | ···················· | ·A·················· | ···················· | ···················· | ···················· | ···················· | |
| P | 8701 hCoV-19/USA/CA-CZB-11166/2020\|EPI_ISL_625524\|2020- | ···················· | ·A·················· | ···················· | ···················· | ···················· | ···················· | |
| P | 8702 hCoV-19/USA/CA-CZB-11167/2020\|EPI_ISL_625525\|2020- | ···················· | ·A·················· | ···················· | ···················· | ···················· | ···················· | |
| P | 8703 hCoV-19/USA/CA-CZB-11169/2020\|EPI_ISL_625526\|2020- | ···················· | ·A·················· | ···················· | ···················· | ···················· | ···················· | |
| P | 8704 hCoV-19/USA/CA-CZB-11170/2020\|EPI_ISL_625527\|2020- | ···················· | ·A·················· | ···················· | ···················· | ···················· | ···················· | |
| P | 8705 hCoV-19/Denmark/DCGC-1774/2020\|EPI_ISL_618200\|2020 | ···················· | ···················· | ···················· | ···················· | ···················· | ···················· | |
| P | 8706 hCoV-19/Denmark/DCGC-1942/2020\|EPI_ISL_618201\|2020 | ···················· | ·A·················· | ···················· | ···················· | ···················· | ···················· | |
| P | 8707 hCoV-19/Denmark/DCGC-1783/2020\|EPI_ISL_618202\|2020 | ···················· | ···················· | ···················· | ···················· | ···················· | ···················· | |
| P | 8708 hCoV-19/Denmark/DCGC-1787/2020\|EPI_ISL_618203\|2020 | ···················· | ···················· | ···················· | ···················· | ···················· | ···················· | |
| P | 8709 hCoV-19/Denmark/DCGC-1791/2020\|EPI_ISL_618204\|2020 | ···················· | ···················· | ···················· | ···················· | ···················· | ···················· | |
| P | 8710 hCoV-19/Denmark/DCGC-1795/2020\|EPI_ISL_618205\|2020 | ···················· | ···················· | ···················· | ···················· | ···················· | ···················· | |
| P | 8711 hCoV-19/Denmark/DCGC-1803/2020\|EPI_ISL_618206\|2020 | ···················· | ···················· | ···················· | ···················· | ···················· | ···················· | |
| P | 8712 hCoV-19/Denmark/DCGC-1805/2020\|EPI_ISL_618207\|2020 | ···················· | ···················· | ···················· | ···················· | ···················· | ···················· | |
| P | 8713 hCoV-19/Denmark/DCGC-1944/2020\|EPI_ISL_618208\|2020 | ···················· | ···················· | ···················· | ···················· | ···················· | ···················· | |
| P | 8714 hCoV-19/Denmark/DCGC-1766/2020\|EPI_ISL_618209\|2020 | ···················· | ···················· | ···················· | ···················· | ···················· | ···················· | |
| P | 8715 hCoV-19/Denmark/DCGC-1771/2020\|EPI_ISL_618210\|2020 | ···················· | ···················· | ···················· | ···················· | ···················· | ···················· | |
| P | 8716 hCoV-19/Denmark/DCGC-1788/2020\|EPI_ISL_618212\|2020 | ···················· | ···················· | ···················· | ···················· | ···················· | ···················· | |
| P | 8717 hCoV-19/Denmark/DCGC-1792/2020\|EPI_ISL_618213\|2020 | ···················· | ···················· | ···················· | ···················· | ···················· | ···················· | |
| P | 8718 hCoV-19/Denmark/DCGC-1796/2020\|EPI_ISL_618214\|2020 | ···················· | ···················· | ···················· | ···················· | ···················· | ···················· | |
| P | 8719 hCoV-19/Denmark/DCGC-1945/2020\|EPI_ISL_618215\|2020 | ···················· | ···················· | ···················· | ···················· | ···················· | ···················· | |
| P | 8720 hCoV-19/Denmark/DCGC-1776/2020\|EPI_ISL_618217\|2020 | ···················· | ···················· | ···················· | ···················· | ···················· | ···················· | |
| P | 8721 hCoV-19/Denmark/DCGC-1779/2020\|EPI_ISL_618218\|2020 | ···················· | ···················· | ···················· | ···················· | ···················· | ···················· | |
| P | 8722 hCoV-19/Denmark/DCGC-1784/2020\|EPI_ISL_618219\|2020 | ···················· | ···················· | ···················· | ···················· | ···················· | ···················· | |
| P | 8723 hCoV-19/Denmark/DCGC-2005/2020\|EPI_ISL_618240\|2020 | ···················· | ···················· | ···················· | ···················· | ···················· | ···················· | |
| P | 8724 hCoV-19/Denmark/DCGC-2010/2020\|EPI_ISL_618241\|2020 | ···················· | ···················· | ···················· | ···················· | ···················· | ···················· | |
| P | 8725 hCoV-19/Denmark/DCGC-2016/2020\|EPI_ISL_618242\|2020 | ···················· | ···················· | ···················· | ···················· | ···················· | ···················· | |
| P | 8726 hCoV-19/Denmark/DCGC-2023/2020\|EPI_ISL_618243\|2020 | ···················· | ···················· | ···················· | ···················· | ···················· | ···················· | |
| P | 8727 hCoV-19/Denmark/DCGC-2076/2020\|EPI_ISL_618244\|2020 | ···················· | ···················· | ···················· | ···················· | ···················· | ···················· | |
| P | 8728 hCoV-19/Denmark/DCGC-2020/2020\|EPI_ISL_618246\|2020 | ···················· | ···················· | ···················· | ···················· | ···················· | ···················· | |
| P | 8729 hCoV-19/Denmark/DCGC-1951/2020\|EPI_ISL_618247\|2020 | ···················· | ···················· | ···················· | ···················· | ···················· | ···················· | |
| P | 8730 hCoV-19/Denmark/DCGC-2025/2020\|EPI_ISL_618248\|2020 | ···················· | ···················· | ···················· | ···················· | ···················· | ···················· | |
| P | 8731 hCoV-19/Denmark/DCGC-2027/2020\|EPI_ISL_618249\|2020 | ···················· | ···················· | ···················· | ···················· | ···················· | ···················· | |
| P | 8732 hCoV-19/Denmark/DCGC-1952/2020\|EPI_ISL_618250\|2020 | ···················· | ···················· | ···················· | ···················· | ···················· | ···················· | |
| P | 8733 hCoV-19/Denmark/DCGC-2003/2020\|EPI_ISL_618251\|2020 | ···················· | ···················· | ···················· | ···················· | ···················· | ···················· | |
| P | 8734 hCoV-19/Denmark/DCGC-2028/2020\|EPI_ISL_618252\|2020 | ···················· | ···················· | ···················· | ···················· | ···················· | ···················· | |
| P | 8735 hCoV-19/Denmark/DCGC-2032/2020\|EPI_ISL_618253\|2020 | ···················· | ···················· | ···················· | ···················· | ···················· | ···················· | |
| P | 8736 hCoV-19/Denmark/DCGC-2034/2020\|EPI_ISL_618254\|2020 | ···················· | ···················· | ···················· | ···················· | ···················· | ···················· | |
| P | 8737 hCoV-19/Denmark/DCGC-2075/2020\|EPI_ISL_618255\|2020 | ···················· | ···················· | ···················· | ···················· | ···················· | ···················· | |
| P | 8738 hCoV-19/Denmark/DCGC-2017/2020\|EPI_ISL_618256\|2020 | ···················· | ···················· | ···················· | ···················· | ···················· | ···················· | |
| P | 8739 hCoV-19/Denmark/DCGC-2029/2020\|EPI_ISL_618257\|2020 | ···················· | ···················· | ···················· | ···················· | ···················· | ···················· | |
| P | 8740 hCoV-19/Denmark/DCGC-2036/2020\|EPI_ISL_618258\|2020 | ···················· | ···················· | ···················· | ···················· | ···················· | ···················· | |
| P | 8741 hCoV-19/Denmark/DCGC-2037/2020\|EPI_ISL_618259\|2020 | ···················· | ···················· | ···················· | ···················· | ···················· | ···················· | |
| P | 8742 hCoV-19/Denmark/DCGC-2038/2020\|EPI_ISL_618260\|2020 | ···················· | ···················· | ···················· | ···················· | ···················· | ···················· | |
| P | 8743 hCoV-19/Denmark/DCGC-2077/2020\|EPI_ISL_618261\|2020 | ···················· | ···················· | ···················· | ···················· | ···················· | ···················· | |
| P | 8744 hCoV-19/Denmark/DCGC-2078/2020\|EPI_ISL_618262\|2020 | ···················· | ···················· | ···················· | ···················· | ···················· | ···················· | |
| P | 8745 hCoV-19/Denmark/DCGC-2079/2020\|EPI_ISL_618263\|2020 | ···················· | ···················· | ···················· | ···················· | ···················· | ···················· | |
| P | 8746 hCoV-19/Denmark/DCGC-2039/2020\|EPI_ISL_618264\|2020 | ···················· | ···················· | ···················· | ···················· | ···················· | ···················· | |
| P | 8747 hCoV-19/Denmark/DCGC-2041/2020\|EPI_ISL_618266\|2020 | ···················· | ···················· | ···················· | ···················· | ···················· | ···················· | |
| P | 8748 hCoV-19/Denmark/DCGC-2042/2020\|EPI_ISL_618267\|2020 | ···················· | ···················· | ···················· | ···················· | ···················· | ···················· | |
| P | 8749 hCoV-19/Denmark/DCGC-2043/2020\|EPI_ISL_618268\|2020 | ···················· | ···················· | ···················· | ···················· | ···················· | ···················· | |
| P | 8750 hCoV-19/Denmark/DCGC-1984/2020\|EPI_ISL_618270\|2020 | ···················· | ···················· | ···················· | ···················· | ···················· | ···················· | |
| P | 8751 hCoV-19/Denmark/DCGC-1985/2020\|EPI_ISL_618271\|2020 | ···················· | ···················· | ···················· | ···················· | ···················· | ···················· | |
| P | 8752 hCoV-19/Denmark/DCGC-1956/2020\|EPI_ISL_618272\|2020 | ···················· | ···················· | ···················· | ···················· | ···················· | ···················· | |
| P | 8753 hCoV-19/Denmark/DCGC-1957/2020\|EPI_ISL_618273\|2020 | ···················· | ·A·················· | ···················· | ···················· | ···················· | ···················· | |
| P | 8754 hCoV-19/Denmark/DCGC-2054/2020\|EPI_ISL_618275\|2020 | ···················· | ···················· | ···················· | ···················· | ···················· | ···················· | |
| P | 8755 hCoV-19/Denmark/DCGC-2055/2020\|EPI_ISL_618276\|2020 | ···················· | ···················· | ···················· | ···················· | ···················· | ···················· | |
| P | 8756 hCoV-19/Denmark/DCGC-1991/2020\|EPI_ISL_618277\|2020 | ···················· | ···················· | ···················· | ···················· | ···················· | ···················· | |

| | | | | | | | |
|---|---|---|---|---|---|---|---|
| P | 8757 hCoV-19/Denmark/DCGC-1959/2020\|EPI_ISL_618278\|2020 | ···················· | ···················· | ···················· | ···················· | ···················· | ···················· |
| P | 8758 hCoV-19/Denmark/DCGC-2056/2020\|EPI_ISL_618279\|2020 | ···················· | ···················· | ···················· | ···················· | ···················· | ···················· |
| P | 8759 hCoV-19/Denmark/DCGC-2059/2020\|EPI_ISL_618300\|2020 | ···················· | ···················· | ···················· | ···················· | ···················· | ···················· |
| P | 8760 hCoV-19/Denmark/DCGC-1971/2020\|EPI_ISL_618301\|2020 | ···················· | ···················· | ···················· | ···················· | ···················· | ···················· |
| P | 8761 hCoV-19/Denmark/DCGC-2060/2020\|EPI_ISL_618302\|2020 | ···················· | ···················· | ···················· | ···················· | ···················· | ···················· |
| P | 8762 hCoV-19/Denmark/DCGC-2061/2020\|EPI_ISL_618303\|2020 | ···················· | ···················· | ···················· | ···················· | ···················· | ···················· |
| P | 8763 hCoV-19/Denmark/DCGC-2062/2020\|EPI_ISL_618304\|2020 | ···················· | ···················· | ···················· | ···················· | ···················· | ···················· |
| P | 8764 hCoV-19/Denmark/DCGC-1972/2020\|EPI_ISL_618305\|2020 | ···················· | ···················· | ···················· | ···················· | ···················· | ···················· |
| P | 8765 hCoV-19/Denmark/DCGC-1973/2020\|EPI_ISL_618306\|2020 | ···················· | ···················· | ···················· | ···················· | ···················· | ···················· |
| P | 8766 hCoV-19/Denmark/DCGC-2063/2020\|EPI_ISL_618307\|2020 | ···················· | ···················· | ···················· | ···················· | ···················· | ···················· |
| P | 8767 hCoV-19/Denmark/DCGC-2064/2020\|EPI_ISL_618308\|2020 | ···················· | ···················· | ···················· | ···················· | ···················· | ···················· |
| P | 8768 hCoV-19/Denmark/DCGC-1974/2020\|EPI_ISL_618309\|2020 | ···················· | ···················· | ···················· | ···················· | ···················· | ···················· |
| P | 8769 hCoV-19/Denmark/DCGC-1975/2020\|EPI_ISL_618310\|2020 | ···················· | ···················· | ···················· | ···················· | ···················· | ···················· |
| P | 8770 hCoV-19/Denmark/DCGC-1976/2020\|EPI_ISL_618311\|2020 | ···················· | ···················· | ···················· | ···················· | ···················· | ···················· |
| P | 8771 hCoV-19/Denmark/DCGC-1977/2020\|EPI_ISL_618312\|2020 | ···················· | ·A·················· | ···················· | ···················· | ···················· | ···················· |
| P | 8772 hCoV-19/Denmark/DCGC-1710/2020\|EPI_ISL_618313\|2020 | ···················· | ···················· | ···················· | ···················· | ···················· | ···················· |
| P | 8773 hCoV-19/Denmark/DCGC-1978/2020\|EPI_ISL_618314\|2020 | ···················· | ···················· | ···················· | ···················· | ···················· | ···················· |
| P | 8774 hCoV-19/Denmark/DCGC-2080/2020\|EPI_ISL_618315\|2020 | ···················· | ···················· | ···················· | ···················· | ···················· | ···················· |
| P | 8775 hCoV-19/Denmark/DCGC-2065/2020\|EPI_ISL_618316\|2020 | ···················· | ·A·················· | ···················· | ···················· | ···················· | ···················· |
| P | 8776 hCoV-19/Denmark/DCGC-2066/2020\|EPI_ISL_618317\|2020 | ···················· | ···G················ | ···················· | ···················· | ···················· | ···················· |
| P | 8777 hCoV-19/Denmark/DCGC-1979/2020\|EPI_ISL_618318\|2020 | ···················· | ···················· | ···················· | ···················· | ···················· | ···················· |
| P | 8778 hCoV-19/Denmark/DCGC-1988/2020\|EPI_ISL_618319\|2020 | ···················· | ···················· | ···················· | ···················· | ···················· | ···················· |
| P | 8779 hCoV-19/Denmark/DCGC-6833/2020\|EPI_ISL_619530\|2020 | ···················· | ···················· | ···················· | ···················· | ···················· | ···················· |
| P | 8780 hCoV-19/Denmark/DCGC-6630/2020\|EPI_ISL_619531\|2020 | ···················· | ···················· | ···················· | ···················· | ···················· | ···················· |
| P | 8781 hCoV-19/Denmark/DCGC-6632/2020\|EPI_ISL_619533\|2020 | ···················· | ···················· | ···················· | ···················· | ···················· | ···················· |
| P | 8782 hCoV-19/Denmark/DCGC-1287/2020\|EPI_ISL_619534\|2020 | ···················· | ···················· | ···················· | ···················· | ···················· | ···················· |
| P | 8783 hCoV-19/Denmark/DCGC-6634/2020\|EPI_ISL_619536\|2020 | ···················· | ···················· | ···················· | ···················· | ···················· | ···················· |
| P | 8784 hCoV-19/Denmark/DCGC-6635/2020\|EPI_ISL_619537\|2020 | ···················· | ···················· | ···················· | ···················· | ···················· | ···················· |
| P | 8785 hCoV-19/Denmark/DCGC-6636/2020\|EPI_ISL_619538\|2020 | ···················· | ···················· | ···················· | ···················· | ···················· | ···················· |
| P | 8786 hCoV-19/Denmark/DCGC-6637/2020\|EPI_ISL_619539\|2020 | ···················· | ···················· | ···················· | ···················· | ···················· | ···················· |
| P | 8787 hCoV-19/Denmark/DCGC-6638/2020\|EPI_ISL_619540\|2020 | ···················· | ···················· | ···················· | ···················· | ···················· | ···················· |
| P | 8788 hCoV-19/Denmark/DCGC-6640/2020\|EPI_ISL_619542\|2020 | ···················· | ···················· | ···················· | ···················· | ···················· | ···················· |
| P | 8789 hCoV-19/Denmark/DCGC-6641/2020\|EPI_ISL_619543\|2020 | ···················· | ···················· | ···················· | ···················· | ···················· | ···················· |
| P | 8790 hCoV-19/Denmark/DCGC-6642/2020\|EPI_ISL_619544\|2020 | ···················· | ·A·················· | ···················· | ···················· | ···················· | ···················· |
| P | 8791 hCoV-19/Denmark/DCGC-6643/2020\|EPI_ISL_619545\|2020 | ···················· | ···················· | ···················· | ···················· | ···················· | ···················· |
| P | 8792 hCoV-19/Denmark/DCGC-6644/2020\|EPI_ISL_619546\|2020 | ···················· | ···················· | ···················· | ···················· | ···················· | ···················· |
| P | 8793 hCoV-19/Denmark/DCGC-6645/2020\|EPI_ISL_619547\|2020 | ···················· | ···················· | ···················· | ···················· | ···················· | ···················· |
| P | 8794 hCoV-19/Denmark/DCGC-6646/2020\|EPI_ISL_619548\|2020 | ···················· | ···················· | ···················· | ···················· | ···················· | ···················· |
| P | 8795 hCoV-19/Denmark/DCGC-6647/2020\|EPI_ISL_619549\|2020 | ···················· | ···················· | ···················· | ···················· | ···················· | ···················· |
| P | 8796 hCoV-19/Denmark/DCGC-6667/2020\|EPI_ISL_619570\|2020 | ···················· | ···················· | ···················· | ···················· | ···················· | ···················· |
| P | 8797 hCoV-19/Denmark/DCGC-6668/2020\|EPI_ISL_619571\|2020 | ···················· | ···················· | ···················· | ···················· | ···················· | ···················· |
| P | 8798 hCoV-19/Denmark/DCGC-6669/2020\|EPI_ISL_619572\|2020 | ···················· | ···················· | ···················· | ···················· | ···················· | ···················· |
| P | 8799 hCoV-19/Denmark/DCGC-6670/2020\|EPI_ISL_619573\|2020 | ···················· | ···················· | ···················· | ···················· | ···················· | ···················· |
| P | 8800 hCoV-19/Denmark/DCGC-6671/2020\|EPI_ISL_619574\|2020 | ···················· | ···················· | ···················· | ···················· | ···················· | ···················· |
| P | 8801 hCoV-19/Denmark/DCGC-6672/2020\|EPI_ISL_619575\|2020 | ···················· | ···················· | ···················· | ···················· | ···················· | ···················· |
| P | 8802 hCoV-19/Denmark/DCGC-6673/2020\|EPI_ISL_619576\|2020 | ···················· | ···················· | ···················· | ···················· | ···················· | ···················· |
| P | 8803 hCoV-19/Denmark/DCGC-6674/2020\|EPI_ISL_619577\|2020 | ···················· | ···················· | ···················· | ···················· | ···················· | ···················· |
| P | 8804 hCoV-19/Denmark/DCGC-6676/2020\|EPI_ISL_619579\|2020 | ···················· | ···················· | ···················· | ···················· | ···················· | ···················· |
| P | 8805 hCoV-19/Denmark/DCGC-6678/2020\|EPI_ISL_619581\|2020 | ···················· | ···················· | ···················· | ···················· | ···················· | ···················· |
| P | 8806 hCoV-19/Denmark/DCGC-6679/2020\|EPI_ISL_619582\|2020 | ···················· | ···················· | ···················· | ···················· | ···················· | ···················· |
| P | 8807 hCoV-19/Denmark/DCGC-6680/2020\|EPI_ISL_619583\|2020 | ···················· | ···················· | ···················· | ···················· | ···················· | ···················· |
| P | 8808 hCoV-19/Denmark/DCGC-6682/2020\|EPI_ISL_619585\|2020 | ···················· | ···················· | ···················· | ···················· | ···················· | ···················· |
| P | 8809 hCoV-19/Denmark/DCGC-6683/2020\|EPI_ISL_619586\|2020 | ···················· | ···················· | ···················· | ···················· | ···················· | ···················· |
| P | 8810 hCoV-19/Denmark/DCGC-6684/2020\|EPI_ISL_619587\|2020 | ···················· | ···················· | ···················· | ···················· | ···················· | ···················· |
| P | 8811 hCoV-19/Denmark/DCGC-6685/2020\|EPI_ISL_619588\|2020 | ···················· | ···················· | ···················· | ···················· | ···················· | ···················· |
| P | 8812 hCoV-19/Denmark/DCGC-6686/2020\|EPI_ISL_619589\|2020 | ···················· | ···················· | ···················· | ···················· | ···················· | ···················· |
| P | 8813 hCoV-19/Denmark/DCGC-6687/2020\|EPI_ISL_619590\|2020 | ···················· | ···················· | ···················· | ···················· | ···················· | ···················· |
| P | 8814 hCoV-19/Denmark/DCGC-6688/2020\|EPI_ISL_619591\|2020 | ···················· | ···················· | ···················· | ···················· | ···················· | ···················· |
| P | 8815 hCoV-19/Denmark/DCGC-6689/2020\|EPI_ISL_619592\|2020 | ···················· | ···················· | ···················· | ···················· | ···················· | ···················· |
| P | 8816 hCoV-19/Denmark/DCGC-6690/2020\|EPI_ISL_619593\|2020 | ···················· | ···················· | ···················· | ···················· | ···················· | ···················· |
| P | 8817 hCoV-19/Denmark/DCGC-6691/2020\|EPI_ISL_619594\|2020 | ···················· | ···················· | ···················· | ···················· | ···················· | ···················· |
| P | 8818 hCoV-19/Denmark/DCGC-6696/2020\|EPI_ISL_619599\|2020 | ···················· | ···················· | ···················· | ···················· | ···················· | ···················· |
| P | 8819 hCoV-19/Denmark/DCGC-6697/2020\|EPI_ISL_619600\|2020 | ···················· | ···················· | ···················· | ···················· | ···················· | ···················· |
| P | 8820 hCoV-19/Denmark/DCGC-6555/2020\|EPI_ISL_619603\|2020 | ···················· | ···················· | ···················· | ···················· | ···················· | ···················· |
| P | 8821 hCoV-19/Denmark/DCGC-6556/2020\|EPI_ISL_619604\|2020 | ···················· | ···················· | ···················· | ···················· | ···················· | ···················· |
| P | 8822 hCoV-19/Denmark/DCGC-6557/2020\|EPI_ISL_619605\|2020 | ···················· | ···················· | ···················· | ···················· | ···················· | ···················· |
| P | 8823 hCoV-19/Denmark/DCGC-6558/2020\|EPI_ISL_619606\|2020 | ···················· | ···················· | ···················· | ···················· | ···················· | ···················· |
| P | 8824 hCoV-19/Denmark/DCGC-6562/2020\|EPI_ISL_619610\|2020 | ···················· | ···················· | ···················· | ···················· | ···················· | ···················· |
| P | 8825 hCoV-19/Denmark/DCGC-6573/2020\|EPI_ISL_619621\|2020 | ···················· | ···················· | ···················· | ···················· | ···················· | ···················· |
| P | 8826 hCoV-19/Denmark/DCGC-6574/2020\|EPI_ISL_619622\|2020 | ···················· | ·A·················· | ···················· | ···················· | ···················· | ···················· |
| P | 8827 hCoV-19/Denmark/DCGC-6575/2020\|EPI_ISL_619623\|2020 | ···················· | ···················· | ···················· | ···················· | ···················· | ···················· |
| P | 8828 hCoV-19/Denmark/DCGC-6576/2020\|EPI_ISL_619624\|2020 | ···················· | ·A·················· | ···················· | ···················· | ···················· | ···················· |
| P | 8829 hCoV-19/Denmark/DCGC-6577/2020\|EPI_ISL_619625\|2020 | ···················· | ···················· | ···················· | ···················· | ···················· | ···················· |
| P | 8830 hCoV-19/Denmark/DCGC-6578/2020\|EPI_ISL_619626\|2020 | ···················· | ···················· | ···················· | ···················· | ···················· | ···················· |
| P | 8831 hCoV-19/Denmark/DCGC-6579/2020\|EPI_ISL_619627\|2020 | ···················· | ···················· | ···················· | ···················· | ···················· | ···················· |
| P | 8832 hCoV-19/Denmark/DCGC-6580/2020\|EPI_ISL_619628\|2020 | ···················· | ···················· | ···················· | ···················· | ···················· | ···················· |
| P | 8833 hCoV-19/Denmark/DCGC-6581/2020\|EPI_ISL_619629\|2020 | ···················· | ···················· | ···················· | ···················· | ···················· | ···················· |
| P | 8834 hCoV-19/Denmark/DCGC-6582/2020\|EPI_ISL_619630\|2020 | ···················· | ···················· | ···················· | ···················· | ···················· | ···················· |
| P | 8835 hCoV-19/Denmark/DCGC-6583/2020\|EPI_ISL_619631\|2020 | ···················· | ···················· | ···················· | ···················· | ···················· | ···················· |
| P | 8836 hCoV-19/Denmark/DCGC-6584/2020\|EPI_ISL_619632\|2020 | ···················· | ···················· | ···················· | ···················· | ···················· | ···················· |

| | | | | | | | | |
|---|---|---|---|---|---|---|---|---|
| P | 8837 | hCoV-19/Denmark/DCGC-6586/2020\|EPI_ISL_619634\|2020 | .................... | .................... | .................... | .................... | .................... | .................... |
| P | 8838 | hCoV-19/Denmark/DCGC-6587/2020\|EPI_ISL_619635\|2020 | .................... | .................... | .................... | .................... | .................... | .................... |
| P | 8839 | hCoV-19/Denmark/DCGC-6588/2020\|EPI_ISL_619636\|2020 | .................... | .................... | .................... | .................... | .................... | .................... |
| P | 8840 | hCoV-19/Denmark/DCGC-6589/2020\|EPI_ISL_619637\|2020 | .................... | .................... | .................... | .................... | .................... | .................... |
| P | 8841 | hCoV-19/Denmark/DCGC-6590/2020\|EPI_ISL_619638\|2020 | .................... | .................... | .................... | .................... | .................... | .................... |
| P | 8842 | hCoV-19/Denmark/DCGC-6591/2020\|EPI_ISL_619639\|2020 | .................... | .................... | .................... | .................... | .................... | .................... |
| P | 8843 | hCoV-19/Denmark/DCGC-6592/2020\|EPI_ISL_619640\|2020 | .................... | .................... | .................... | .................... | .................... | .................... |
| P | 8844 | hCoV-19/Denmark/DCGC-6597/2020\|EPI_ISL_619645\|2020 | .................... | .................... | .................... | .................... | .................... | .................... |
| P | 8845 | hCoV-19/Denmark/DCGC-6598/2020\|EPI_ISL_619646\|2020 | .................... | .................... | .................... | .................... | .................... | .................... |
| P | 8846 | hCoV-19/Denmark/DCGC-6599/2020\|EPI_ISL_619647\|2020 | .................... | .................... | .................... | .................... | .................... | .................... |
| P | 8847 | hCoV-19/Denmark/DCGC-6600/2020\|EPI_ISL_619648\|2020 | .................... | .A.................. | .................... | .................... | .................... | ...A................ |
| P | 8848 | hCoV-19/Denmark/DCGC-6601/2020\|EPI_ISL_619649\|2020 | .................... | .................... | .................... | .................... | .................... | .................... |
| P | 8849 | hCoV-19/Denmark/DCGC-4422/2020\|EPI_ISL_621843\|2020 | .................... | .................... | .................... | .................... | .................... | .................... |
| P | 8850 | hCoV-19/Denmark/DCGC-4423/2020\|EPI_ISL_621844\|2020 | .................... | .................... | .................... | .................... | .................... | .................... |
| P | 8851 | hCoV-19/Denmark/DCGC-4424/2020\|EPI_ISL_621845\|2020 | .................... | .................... | .................... | .................... | .................... | .................... |
| P | 8852 | hCoV-19/Denmark/DCGC-4425/2020\|EPI_ISL_621846\|2020 | .................... | .................... | .................... | .................... | .................... | .................... |
| P | 8853 | hCoV-19/Denmark/DCGC-4426/2020\|EPI_ISL_621847\|2020 | .................... | .................... | .................... | .................... | .................... | .................... |
| P | 8854 | hCoV-19/Denmark/DCGC-4427/2020\|EPI_ISL_621848\|2020 | .................... | .................... | .................... | .................... | .................... | .................... |
| P | 8855 | hCoV-19/Denmark/DCGC-4428/2020\|EPI_ISL_621849\|2020 | .................... | .................... | .................... | .................... | .................... | .................... |
| P | 8856 | hCoV-19/Denmark/DCGC-4434/2020\|EPI_ISL_621855\|2020 | .................... | .................... | .................... | .................... | .................... | .................... |
| P | 8857 | hCoV-19/Denmark/DCGC-4436/2020\|EPI_ISL_621857\|2020 | .................... | .................... | .................... | .................... | .................... | .................... |
| P | 8858 | hCoV-19/Denmark/DCGC-4437/2020\|EPI_ISL_621858\|2020 | .................... | .................... | .................... | .................... | .................... | .................... |
| P | 8859 | hCoV-19/Denmark/DCGC-4438/2020\|EPI_ISL_621859\|2020 | .................... | .................... | .................... | .................... | .................... | .................... |
| P | 8860 | hCoV-19/Denmark/DCGC-4439/2020\|EPI_ISL_621860\|2020 | .................... | .................... | .................... | .................... | .................... | .................... |
| P | 8861 | hCoV-19/Denmark/DCGC-4440/2020\|EPI_ISL_621861\|2020 | .................... | .................... | .................... | .................... | .................... | .................... |
| P | 8862 | hCoV-19/Denmark/DCGC-4441/2020\|EPI_ISL_621862\|2020 | .................... | .................... | .................... | .................... | .................... | .................... |
| P | 8863 | hCoV-19/Denmark/DCGC-4442/2020\|EPI_ISL_621863\|2020 | .................... | .................... | .................... | .................... | .................... | .................... |
| P | 8864 | hCoV-19/Denmark/DCGC-4443/2020\|EPI_ISL_621864\|2020 | .................... | .................... | .................... | .................... | .................... | .................... |
| P | 8865 | hCoV-19/Denmark/DCGC-4655/2020\|EPI_ISL_621887\|2020 | .................... | .................... | .................... | .................... | .................... | .................... |
| P | 8866 | hCoV-19/Denmark/DCGC-4656/2020\|EPI_ISL_621888\|2020 | .................... | .................... | .................... | .................... | .................... | .................... |
| P | 8867 | hCoV-19/Denmark/DCGC-4658/2020\|EPI_ISL_621890\|2020 | .................... | .................... | .................... | .................... | .................... | .................... |
| P | 8868 | hCoV-19/Denmark/DCGC-4659/2020\|EPI_ISL_621891\|2020 | .................... | .................... | .................... | .................... | .................... | .................... |
| P | 8869 | hCoV-19/Denmark/DCGC-4660/2020\|EPI_ISL_621892\|2020 | .................... | .................... | .................... | .................... | .................... | .................... |
| P | 8870 | hCoV-19/Denmark/DCGC-4661/2020\|EPI_ISL_621893\|2020 | .................... | .................... | .................... | .................... | .................... | .................... |
| P | 8871 | hCoV-19/Denmark/DCGC-4662/2020\|EPI_ISL_621894\|2020 | .................... | .................... | .................... | .................... | .................... | .................... |
| P | 8872 | hCoV-19/Denmark/DCGC-4663/2020\|EPI_ISL_621895\|2020 | .................... | .................... | .................... | .................... | .................... | .................... |
| P | 8873 | hCoV-19/Denmark/DCGC-4664/2020\|EPI_ISL_621896\|2020 | .................... | .................... | .................... | .................... | .................... | .................... |
| P | 8874 | hCoV-19/Denmark/DCGC-4665/2020\|EPI_ISL_621897\|2020 | .................... | .................... | .................... | .................... | .................... | .................... |
| P | 8875 | hCoV-19/Denmark/DCGC-4667/2020\|EPI_ISL_621899\|2020 | .................... | .................... | .................... | .................... | .................... | .................... |
| P | 8876 | hCoV-19/Denmark/DCGC-4668/2020\|EPI_ISL_621900\|2020 | .................... | .................... | .................... | .................... | .................... | .................... |
| P | 8877 | hCoV-19/Denmark/DCGC-4669/2020\|EPI_ISL_621901\|2020 | .................... | .................... | .................... | .................... | .................... | .................... |
| P | 8878 | hCoV-19/Denmark/DCGC-4670/2020\|EPI_ISL_621902\|2020 | .................... | .................... | .................... | .................... | .................... | .................... |
| P | 8879 | hCoV-19/Denmark/DCGC-4671/2020\|EPI_ISL_621903\|2020 | .................... | .................... | .................... | .................... | .................... | .................... |
| P | 8880 | hCoV-19/Denmark/DCGC-4672/2020\|EPI_ISL_621904\|2020 | .................... | .................... | .................... | .................... | .................... | .................... |
| P | 8881 | hCoV-19/Denmark/DCGC-4673/2020\|EPI_ISL_621905\|2020 | .................... | .................... | .................... | .................... | .................... | .................... |
| P | 8882 | hCoV-19/Denmark/DCGC-4674/2020\|EPI_ISL_621906\|2020 | .................... | .................... | .................... | .................... | .................... | .................... |
| P | 8883 | hCoV-19/Denmark/DCGC-4675/2020\|EPI_ISL_621907\|2020 | .................... | .................... | .................... | .................... | .................... | .................... |
| P | 8884 | hCoV-19/Denmark/DCGC-4676/2020\|EPI_ISL_621908\|2020 | .................... | .................... | .................... | .................... | .................... | .................... |
| P | 8885 | hCoV-19/Denmark/DCGC-4677/2020\|EPI_ISL_621909\|2020 | .................... | .................... | .................... | .................... | .................... | .................... |
| P | 8886 | hCoV-19/Denmark/DCGC-4678/2020\|EPI_ISL_621910\|2020 | .................... | .................... | .................... | .................... | .................... | .................... |
| P | 8887 | hCoV-19/Denmark/DCGC-4679/2020\|EPI_ISL_621911\|2020 | .................... | .................... | .................... | .................... | .................... | .................... |
| P | 8888 | hCoV-19/Denmark/DCGC-4680/2020\|EPI_ISL_621912\|2020 | .................... | .................... | .................... | .................... | .................... | .................... |
| P | 8889 | hCoV-19/Denmark/DCGC-4681/2020\|EPI_ISL_621913\|2020 | .................... | .................... | .................... | .................... | .................... | .................... |
| P | 8890 | hCoV-19/Denmark/DCGC-4682/2020\|EPI_ISL_621914\|2020 | .................... | .................... | .................... | .................... | .................... | .................... |
| P | 8891 | hCoV-19/Denmark/DCGC-4683/2020\|EPI_ISL_621915\|2020 | .................... | .................... | .................... | .................... | .................... | .................... |
| P | 8892 | hCoV-19/Denmark/DCGC-4684/2020\|EPI_ISL_621916\|2020 | .................... | .................... | .................... | .................... | .................... | .................... |
| P | 8893 | hCoV-19/Denmark/DCGC-4686/2020\|EPI_ISL_621918\|2020 | .................... | .................... | .................... | .................... | .................... | .................... |
| P | 8894 | hCoV-19/Denmark/DCGC-4687/2020\|EPI_ISL_621919\|2020 | .................... | .................... | .................... | .................... | .................... | .................... |
| P | 8895 | hCoV-19/Denmark/DCGC-4688/2020\|EPI_ISL_621920\|2020 | .................... | .................... | .................... | .................... | .................... | .................... |
| P | 8896 | hCoV-19/Denmark/DCGC-4689/2020\|EPI_ISL_621921\|2020 | .................... | .................... | .................... | .................... | .................... | .................... |
| P | 8897 | hCoV-19/Denmark/DCGC-4690/2020\|EPI_ISL_621922\|2020 | .................... | .................... | .................... | .................... | .................... | .................... |
| P | 8898 | hCoV-19/Denmark/DCGC-4691/2020\|EPI_ISL_621923\|2020 | .................... | .................... | .................... | .................... | .................... | .................... |
| P | 8899 | hCoV-19/Denmark/DCGC-4692/2020\|EPI_ISL_621924\|2020 | .................... | .................... | .................... | .................... | .................... | .................... |
| P | 8900 | hCoV-19/Denmark/DCGC-4693/2020\|EPI_ISL_621925\|2020 | .................... | .................... | .................... | .................... | .................... | .................... |
| P | 8901 | hCoV-19/Denmark/DCGC-4694/2020\|EPI_ISL_621926\|2020 | .................... | .A.................. | .................... | .................... | .................... | .................... |
| P | 8902 | hCoV-19/Denmark/DCGC-4695/2020\|EPI_ISL_621927\|2020 | .................... | .................... | .................... | .................... | .................... | .................... |
| P | 8903 | hCoV-19/Denmark/DCGC-4696/2020\|EPI_ISL_621928\|2020 | .................... | .................... | .................... | .................... | .................... | .................... |
| P | 8904 | hCoV-19/Denmark/DCGC-4710/2020\|EPI_ISL_621941\|2020 | .................... | .................... | .................... | .................... | .................... | .................... |
| P | 8905 | hCoV-19/Denmark/DCGC-4122/2020\|EPI_ISL_621942\|2020 | .................... | .................... | .................... | .................... | .................... | .................... |
| P | 8906 | hCoV-19/Denmark/DCGC-4124/2020\|EPI_ISL_621944\|2020 | .................... | .................... | .................... | .................... | .................... | .................... |
| P | 8907 | hCoV-19/Denmark/DCGC-4125/2020\|EPI_ISL_621945\|2020 | .................... | .................... | .................... | .................... | .................... | .................... |
| P | 8908 | hCoV-19/Denmark/DCGC-4127/2020\|EPI_ISL_621947\|2020 | .................... | .................... | .................... | .................... | .................... | .................... |
| P | 8909 | hCoV-19/Denmark/DCGC-4128/2020\|EPI_ISL_621948\|2020 | .................... | .................... | .................... | .................... | .................... | .................... |
| P | 8910 | hCoV-19/Denmark/DCGC-4129/2020\|EPI_ISL_621949\|2020 | .................... | .................... | .................... | .................... | .................... | .................... |
| P | 8911 | hCoV-19/Denmark/DCGC-4130/2020\|EPI_ISL_621950\|2020 | .................... | .................... | .................... | .................... | .................... | .................... |
| P | 8912 | hCoV-19/Denmark/DCGC-4131/2020\|EPI_ISL_621951\|2020 | .................... | .................... | .................... | .................... | .................... | .................... |
| P | 8913 | hCoV-19/Denmark/DCGC-4132/2020\|EPI_ISL_621952\|2020 | .................... | .................... | .................... | .................... | .................... | .................... |
| P | 8914 | hCoV-19/Denmark/DCGC-4133/2020\|EPI_ISL_621953\|2020 | .................... | .................... | .................... | .................... | .................... | .................... |
| P | 8915 | hCoV-19/Denmark/DCGC-4134/2020\|EPI_ISL_621954\|2020 | .................... | .................... | .................... | .................... | .................... | .................... |
| P | 8916 | hCoV-19/Denmark/DCGC-4135/2020\|EPI_ISL_621955\|2020 | .................... | .................... | .................... | .................... | .................... | .................... |

| | | | | | | | |
|---|---|---|---|---|---|---|---|
| P | 8917 hCoV-19/Denmark/DCGC-4136/2020\|EPI_ISL_621956\|2020 | ···················· | ···················· | ···················· | ···················· | ···················· | ···················· |
| P | 8918 hCoV-19/Denmark/DCGC-4139/2020\|EPI_ISL_621959\|2020 | ···················· | ···················· | ···················· | ···················· | ···················· | ···················· |
| P | 8919 hCoV-19/Denmark/DCGC-4140/2020\|EPI_ISL_621960\|2020 | ···················· | ···················· | ···················· | ···················· | ···················· | ···················· |
| P | 8920 hCoV-19/Denmark/DCGC-4141/2020\|EPI_ISL_621961\|2020 | ···················· | ···················· | ···················· | ···················· | ···················· | ···················· |
| P | 8921 hCoV-19/Denmark/DCGC-4142/2020\|EPI_ISL_621962\|2020 | ···················· | ···················· | ···················· | ···················· | ···················· | ···················· |
| P | 8922 hCoV-19/Denmark/DCGC-4143/2020\|EPI_ISL_621963\|2020 | ···················· | ···················· | ···················· | ···················· | ···················· | ···················· |
| P | 8923 hCoV-19/England/ALDP-AA079A/2020\|EPI_ISL_624160\|20 | ···················· | ···················· | ···················· | ···················· | ···················· | ···················· |
| P | 8924 hCoV-19/England/ALDP-AA191B/2020\|EPI_ISL_624161\|20 | ···················· | ···················· | ···················· | ···················· | ···················· | ···················· |
| P | 8925 hCoV-19/England/ALDP-AA3348/2020\|EPI_ISL_624162\|20 | ···················· | ···················· | ···················· | ···················· | ···················· | ···················· |
| P | 8926 hCoV-19/Scotland/QEUH-AADCCB/2020\|EPI_ISL_624164\|2 | ···················· | ···················· | ···················· | ···················· | ···················· | ···················· |
| P | 8927 hCoV-19/England/QEUH-AADD22/2020\|EPI_ISL_624165\|20 | ···················· | ···················· | ···················· | ···················· | ···················· | ···················· |
| P | 8928 hCoV-19/England/ALDP-AA130E/2020\|EPI_ISL_624167\|20 | ···················· | ···················· | ···················· | ···················· | ···················· | ···················· |
| P | 8929 hCoV-19/England/ALDP-AA074F/2020\|EPI_ISL_624168\|20 | ···················· | ·A·················· | ···················· | ···················· | ···················· | ···················· |
| P | 8930 hCoV-19/England/ALDP-AA357F/2020\|EPI_ISL_624169\|20 | ···················· | ···················· | ···················· | ···················· | ···················· | ···················· |
| P | 8931 hCoV-19/England/ALDP-AA373D/2020\|EPI_ISL_624170\|20 | ···················· | ···················· | ···················· | ···················· | ···················· | ···················· |
| P | 8932 hCoV-19/England/ALDP-AA2F90/2020\|EPI_ISL_624171\|20 | ···················· | ···················· | ···················· | ···················· | ···················· | ···················· |
| P | 8933 hCoV-19/England/ALDP-AA372E/2020\|EPI_ISL_624172\|20 | ···················· | ···················· | ···················· | ···················· | ···················· | ···················· |
| P | 8934 hCoV-19/England/ALDP-AA14DE/2020\|EPI_ISL_624175\|20 | ···················· | ···················· | ···················· | ···················· | ···················· | ···················· |
| P | 8935 hCoV-19/Scotland/QEUH-AADCAD/2020\|EPI_ISL_624176\|2 | ···················· | ···················· | ···················· | ···················· | ···················· | ···················· |
| P | 8936 hCoV-19/England/ALDP-AA1948/2020\|EPI_ISL_624177\|20 | ···················· | ···················· | ···················· | ···················· | ···················· | ···················· |
| P | 8937 hCoV-19/England/ALDP-AA5179/2020\|EPI_ISL_624178\|20 | ···················· | ···················· | ···················· | ···················· | ···················· | ···················· |
| P | 8938 hCoV-19/England/ALDP-AA190C/2020\|EPI_ISL_624179\|20 | ···················· | ···················· | ···················· | ···················· | ···················· | ···················· |
| P | 8939 hCoV-19/Scotland/QEUH-AAEEE2/2020\|EPI_ISL_624181\|2 | ···················· | ···················· | ···················· | ···················· | ···················· | ···················· |
| P | 8940 hCoV-19/England/ALDP-AA1957/2020\|EPI_ISL_624182\|20 | ···················· | ···················· | ···················· | ···················· | ···················· | ···················· |
| P | 8941 hCoV-19/England/ALDP-AA3199/2020\|EPI_ISL_624183\|20 | ···················· | ···················· | ···················· | ···················· | ···················· | ···················· |
| P | 8942 hCoV-19/England/ALDP-AA2F72/2020\|EPI_ISL_624184\|20 | ···················· | ···················· | ···················· | ···················· | ···················· | ···················· |
| P | 8943 hCoV-19/England/ALDP-AA169C/2020\|EPI_ISL_624185\|20 | ···················· | ···················· | ···················· | ···················· | ···················· | ···················· |
| P | 8944 hCoV-19/England/QEUH-AAE486/2020\|EPI_ISL_624188\|20 | ···················· | ···················· | ···················· | ···················· | ···················· | ···················· |
| P | 8945 hCoV-19/Scotland/QEUH-AAD311/2020\|EPI_ISL_624189\|2 | ···················· | ···················· | ···················· | ···················· | ···················· | ···················· |
| P | 8946 hCoV-19/England/ALDP-AA31A8/2020\|EPI_ISL_624190\|20 | ···················· | ···················· | ···················· | ···················· | ···················· | ···················· |
| P | 8947 hCoV-19/England/ALDP-AA2D78/2020\|EPI_ISL_624191\|20 | ···················· | ···················· | ···················· | ···················· | ···················· | ···················· |
| P | 8948 hCoV-19/England/ALDP-AA415C/2020\|EPI_ISL_624193\|20 | ···················· | ···················· | ···················· | ···················· | ···················· | ···················· |
| P | 8949 hCoV-19/England/ALDP-AA2F27/2020\|EPI_ISL_624194\|20 | ···················· | ···················· | ···················· | ···················· | ···················· | ···················· |
| P | 8950 hCoV-19/England/ALDP-AA30BA/2020\|EPI_ISL_624195\|20 | ···················· | ···················· | ···················· | ···················· | ···················· | ···················· |
| P | 8951 hCoV-19/England/ALDP-AA2D96/2020\|EPI_ISL_624198\|20 | ···················· | ···················· | ···················· | ···················· | ···················· | ···················· |
| P | 8952 hCoV-19/USA/CA-CZB-11678/2020\|EPI_ISL_625490\|2020- | ···················· | ·A·················· | ···················· | ···················· | ···················· | ···················· |
| P | 8953 hCoV-19/USA/CA-CZB-11681/2020\|EPI_ISL_625492\|2020- | ···················· | ·A·················· | ···················· | ···················· | ···················· | ···················· |
| P | 8954 hCoV-19/USA/CA-CZB-11682/2020\|EPI_ISL_625493\|2020- | ···················· | ·A·················· | ···················· | ···················· | ···················· | ···················· |
| P | 8955 hCoV-19/USA/CA-CZB-11683/2020\|EPI_ISL_625494\|2020- | ···················· | ·A·················· | ···················· | ···················· | ···················· | ···················· |
| P | 8956 hCoV-19/USA/CA-CZB-11686/2020\|EPI_ISL_625495\|2020- | ···················· | ·A·················· | ···················· | ···················· | ···················· | ···················· |
| P | 8957 hCoV-19/USA/CA-CZB-11690/2020\|EPI_ISL_625496\|2020- | ···················· | ···················· | ···················· | ···················· | ···················· | ···················· |
| P | 8958 hCoV-19/USA/CA-CZB-11691/2020\|EPI_ISL_625497\|2020- | ···················· | ···················· | ···················· | ···················· | ···················· | ···················· |
| P | 8959 hCoV-19/USA/CA-CZB-11692/2020\|EPI_ISL_625498\|2020- | ···················· | ·A·················· | ···················· | ···················· | ···················· | ···················· |
| P | 8960 hCoV-19/USA/CA-CZB-11693/2020\|EPI_ISL_625499\|2020- | ···················· | ·A·················· | ···················· | ···················· | ···················· | ···················· |
| P | 8961 hCoV-19/Denmark/DCGC-1169/2020\|EPI_ISL_614639\|2020 | ···················· | ···················· | ···················· | ···················· | ···················· | ···················· |
| P | 8962 hCoV-19/Denmark/DCGC-1139/2020\|EPI_ISL_614640\|2020 | ···················· | ···················· | ···················· | ···················· | ···················· | ···················· |
| P | 8963 hCoV-19/Denmark/DCGC-1156/2020\|EPI_ISL_614641\|2020 | ···················· | ···················· | ···················· | ···················· | ···················· | ···················· |
| P | 8964 hCoV-19/Denmark/DCGC-1162/2020\|EPI_ISL_614642\|2020 | ···················· | ···················· | ···················· | ···················· | ···················· | ···················· |
| P | 8965 hCoV-19/Denmark/DCGC-1182/2020\|EPI_ISL_614643\|2020 | ···················· | ···················· | ···················· | ···················· | ···················· | ···················· |
| P | 8966 hCoV-19/Denmark/DCGC-1142/2020\|EPI_ISL_614644\|2020 | ···················· | ···················· | ···················· | ···················· | ···················· | ···················· |
| P | 8967 hCoV-19/Denmark/DCGC-1151/2020\|EPI_ISL_614645\|2020 | ···················· | ···················· | ···················· | ···················· | ···················· | ···················· |
| P | 8968 hCoV-19/Denmark/DCGC-1177/2020\|EPI_ISL_614646\|2020 | ···················· | ···················· | ···················· | ···················· | ···················· | ···················· |
| P | 8969 hCoV-19/Denmark/DCGC-1152/2020\|EPI_ISL_614647\|2020 | ···················· | ···················· | ···················· | ···················· | ···················· | ···················· |
| P | 8970 hCoV-19/Denmark/DCGC-1183/2020\|EPI_ISL_614648\|2020 | ···················· | ···················· | ···················· | ···················· | ···················· | ···················· |
| P | 8971 hCoV-19/Denmark/DCGC-1049/2020\|EPI_ISL_614680\|2020 | ···················· | ···················· | ···················· | ···················· | ···················· | ···················· |
| P | 8972 hCoV-19/Denmark/DCGC-1050/2020\|EPI_ISL_614681\|2020 | ···················· | ···················· | ···················· | ···················· | ···················· | ···················· |
| P | 8973 hCoV-19/Denmark/DCGC-1051/2020\|EPI_ISL_614682\|2020 | ···················· | ···················· | ···················· | ···················· | ···················· | ···················· |
| P | 8974 hCoV-19/Denmark/DCGC-1052/2020\|EPI_ISL_614683\|2020 | ···················· | ···················· | ···················· | ···················· | ···················· | ···················· |
| P | 8975 hCoV-19/Denmark/DCGC-1053/2020\|EPI_ISL_614684\|2020 | ···················· | ···················· | ···················· | ···················· | ···················· | ···················· |
| P | 8976 hCoV-19/Denmark/DCGC-1054/2020\|EPI_ISL_614685\|2020 | ···················· | ···················· | ···················· | ···················· | ···················· | ···················· |
| P | 8977 hCoV-19/Denmark/DCGC-1055/2020\|EPI_ISL_614686\|2020 | ···················· | ···················· | ···················· | ···················· | ···················· | ···················· |
| P | 8978 hCoV-19/Denmark/DCGC-1056/2020\|EPI_ISL_614687\|2020 | ···················· | ···················· | ···················· | ···················· | ···················· | ···················· |
| P | 8979 hCoV-19/Denmark/DCGC-1057/2020\|EPI_ISL_614688\|2020 | ···················· | ···················· | ···················· | ···················· | ···················· | ···················· |
| P | 8980 hCoV-19/Denmark/DCGC-1058/2020\|EPI_ISL_614689\|2020 | ···················· | ···················· | ···················· | ···················· | ···················· | ···················· |
| P | 8981 hCoV-19/Denmark/DCGC-1059/2020\|EPI_ISL_614690\|2020 | ···················· | ···················· | ···················· | ···················· | ···················· | ···················· |
| P | 8982 hCoV-19/Denmark/DCGC-1060/2020\|EPI_ISL_614691\|2020 | ···················· | ···················· | ···················· | ···················· | ···················· | ···················· |
| P | 8983 hCoV-19/Denmark/DCGC-1061/2020\|EPI_ISL_614692\|2020 | ···················· | ···················· | ···················· | ···················· | ···················· | ···················· |
| P | 8984 hCoV-19/Denmark/DCGC-1062/2020\|EPI_ISL_614693\|2020 | ···················· | ···················· | ···················· | ···················· | ···················· | ···················· |
| P | 8985 hCoV-19/Denmark/DCGC-1063/2020\|EPI_ISL_614694\|2020 | ···················· | ···················· | ···················· | ···················· | ···················· | ···················· |
| P | 8986 hCoV-19/Denmark/DCGC-935/2020\|EPI_ISL_614695\|2020- | ···················· | ···················· | ···················· | ···················· | ···················· | ···················· |
| P | 8987 hCoV-19/Denmark/DCGC-939/2020\|EPI_ISL_614696\|2020- | ···················· | ···················· | ···················· | ···················· | ···················· | ···················· |
| P | 8988 hCoV-19/Denmark/DCGC-793/2020\|EPI_ISL_614697\|2020- | ···················· | ···················· | ···················· | ···················· | ···················· | ···················· |
| P | 8989 hCoV-19/Denmark/DCGC-797/2020\|EPI_ISL_614698\|2020- | ···················· | ···················· | ···················· | ···················· | ···················· | ···················· |
| P | 8990 hCoV-19/Denmark/DCGC-805/2020\|EPI_ISL_614699\|2020- | ···················· | ···················· | ···················· | ···················· | ···················· | ···················· |
| P | 8991 hCoV-19/Denmark/DCGC-809/2020\|EPI_ISL_614700\|2020- | ···················· | ···················· | ···················· | ···················· | ···················· | ···················· |
| P | 8992 hCoV-19/Denmark/DCGC-814/2020\|EPI_ISL_614701\|2020- | ···················· | ···················· | ···················· | ···················· | ···················· | ···················· |
| P | 8993 hCoV-19/Denmark/DCGC-820/2020\|EPI_ISL_614702\|2020- | ···················· | ···················· | ···················· | ···················· | ···················· | ···················· |
| P | 8994 hCoV-19/Denmark/DCGC-834/2020\|EPI_ISL_614703\|2020- | ···················· | ···················· | ···················· | ···················· | ···················· | ···················· |
| P | 8995 hCoV-19/Denmark/DCGC-838/2020\|EPI_ISL_614704\|2020- | ···················· | ···················· | ···················· | ···················· | ···················· | ···················· |
| P | 8996 hCoV-19/Denmark/DCGC-794/2020\|EPI_ISL_614705\|2020- | ···················· | ···················· | ···················· | ···················· | ···················· | ···················· |

| | | | | | | | | |
|---|---|---|---|---|---|---|---|---|
| P | 8997 | hCoV-19/Denmark/DCGC-798/2020\|EPI_ISL_614706\|2020- | .................... | .................... | .................... | .................... | .................... | .................... |
| P | 8998 | hCoV-19/Denmark/DCGC-806/2020\|EPI_ISL_614707\|2020- | .................... | .................... | .................... | .................... | .................... | .................... |
| P | 8999 | hCoV-19/Denmark/DCGC-815/2020\|EPI_ISL_614708\|2020- | .................... | .................... | .................... | .................... | .................... | .................... |
| P | 9000 | hCoV-19/Denmark/DCGC-821/2020\|EPI_ISL_614709\|2020- | .................... | .................... | .................... | .................... | .................... | .................... |
| P | 9001 | hCoV-19/Denmark/DCGC-824/2020\|EPI_ISL_614710\|2020- | .................... | .................... | .................... | .................... | .................... | .................... |
| P | 9002 | hCoV-19/Denmark/DCGC-795/2020\|EPI_ISL_614711\|2020- | .................... | .................... | .................... | .................... | .................... | .................... |
| P | 9003 | hCoV-19/Denmark/DCGC-799/2020\|EPI_ISL_614712\|2020- | .................... | .................... | .................... | .................... | .................... | .................... |
| P | 9004 | hCoV-19/Denmark/DCGC-912/2020\|EPI_ISL_614713\|2020- | .................... | .................... | .................... | .................... | .................... | .................... |
| P | 9005 | hCoV-19/Denmark/DCGC-810/2020\|EPI_ISL_614714\|2020- | .................... | .................... | .................... | .................... | .................... | .................... |
| P | 9006 | hCoV-19/Denmark/DCGC-816/2020\|EPI_ISL_614715\|2020- | .................... | .................... | .................... | .................... | .................... | .................... |
| P | 9007 | hCoV-19/Denmark/DCGC-840/2020\|EPI_ISL_614716\|2020- | .................... | .................... | .................... | .................... | .................... | .................... |
| P | 9008 | hCoV-19/Denmark/DCGC-847/2020\|EPI_ISL_614717\|2020- | .................... | .................... | .................... | .................... | .................... | .................... |
| P | 9009 | hCoV-19/Denmark/DCGC-796/2020\|EPI_ISL_614718\|2020- | .................... | .................... | .................... | .................... | .................... | .................... |
| P | 9010 | hCoV-19/Denmark/DCGC-800/2020\|EPI_ISL_614719\|2020- | .................... | .................... | .................... | .................... | .................... | .................... |
| P | 9011 | hCoV-19/Denmark/DCGC-811/2020\|EPI_ISL_614720\|2020- | .................... | .................... | .................... | .................... | .................... | .................... |
| P | 9012 | hCoV-19/Denmark/DCGC-825/2020\|EPI_ISL_614721\|2020- | .................... | .................... | .................... | .................... | .................... | .................... |
| P | 9013 | hCoV-19/Denmark/DCGC-830/2020\|EPI_ISL_614722\|2020- | .................... | .................... | .................... | .................... | .................... | .................... |
| P | 9014 | hCoV-19/Denmark/DCGC-841/2020\|EPI_ISL_614723\|2020- | .................... | .................... | .................... | .................... | .................... | .................... |
| P | 9015 | hCoV-19/Denmark/DCGC-848/2020\|EPI_ISL_614724\|2020- | .................... | .................... | .................... | .................... | .................... | .................... |
| P | 9016 | hCoV-19/Denmark/DCGC-801/2020\|EPI_ISL_614725\|2020- | .................... | .................... | .................... | .................... | .................... | .................... |
| P | 9017 | hCoV-19/Denmark/DCGC-812/2020\|EPI_ISL_614726\|2020- | .................... | .................... | .................... | .................... | .................... | .................... |
| P | 9018 | hCoV-19/Denmark/DCGC-822/2020\|EPI_ISL_614727\|2020- | .................... | .................... | .................... | .................... | .................... | .................... |
| P | 9019 | hCoV-19/Denmark/DCGC-826/2020\|EPI_ISL_614728\|2020- | .................... | .................... | .................... | .................... | .................... | .................... |
| P | 9020 | hCoV-19/Denmark/DCGC-831/2020\|EPI_ISL_614729\|2020- | .................... | .................... | .................... | .................... | .................... | .................... |
| P | 9021 | hCoV-19/Denmark/DCGC-842/2020\|EPI_ISL_614730\|2020- | .................... | .................... | .................... | .................... | .................... | .................... |
| P | 9022 | hCoV-19/Denmark/DCGC-802/2020\|EPI_ISL_614731\|2020- | .................... | .................... | .................... | .................... | .................... | .................... |
| P | 9023 | hCoV-19/Denmark/DCGC-807/2020\|EPI_ISL_614732\|2020- | .................... | .................... | .................... | .................... | .................... | .................... |
| P | 9024 | hCoV-19/Denmark/DCGC-913/2020\|EPI_ISL_614733\|2020- | .................... | .................... | .................... | .................... | .................... | .................... |
| P | 9025 | hCoV-19/Denmark/DCGC-813/2020\|EPI_ISL_614734\|2020- | .................... | .................... | .................... | .................... | .................... | .................... |
| P | 9026 | hCoV-19/Denmark/DCGC-818/2020\|EPI_ISL_614735\|2020- | .................... | .................... | .................... | .................... | .................... | .................... |
| P | 9027 | hCoV-19/Denmark/DCGC-827/2020\|EPI_ISL_614736\|2020- | .................... | .................... | .................... | .................... | .................... | .................... |
| P | 9028 | hCoV-19/Denmark/DCGC-835/2020\|EPI_ISL_614737\|2020- | .................... | .................... | .................... | .................... | .................... | .................... |
| P | 9029 | hCoV-19/Denmark/DCGC-803/2020\|EPI_ISL_614738\|2020- | .................... | .................... | .................... | .................... | .................... | .................... |
| P | 9030 | hCoV-19/Denmark/DCGC-819/2020\|EPI_ISL_614739\|2020- | .................... | .................... | .................... | .................... | .................... | .................... |
| P | 9031 | hCoV-19/Denmark/DCGC-828/2020\|EPI_ISL_614740\|2020- | .................... | .................... | .................... | .................... | .................... | .................... |
| P | 9032 | hCoV-19/Denmark/DCGC-832/2020\|EPI_ISL_614741\|2020- | .................... | .................... | .................... | .................... | .................... | .................... |
| P | 9033 | hCoV-19/Denmark/DCGC-844/2020\|EPI_ISL_614742\|2020- | .................... | .................... | .................... | .................... | .................... | .................... |
| P | 9034 | hCoV-19/Denmark/DCGC-804/2020\|EPI_ISL_614743\|2020- | .................... | .................... | .................... | .................... | .................... | .................... |
| P | 9035 | hCoV-19/Denmark/DCGC-808/2020\|EPI_ISL_614744\|2020- | .................... | .................... | .................... | .................... | .................... | .................... |
| P | 9036 | hCoV-19/Denmark/DCGC-823/2020\|EPI_ISL_614745\|2020- | .................... | .................... | .................... | .................... | .................... | .................... |
| P | 9037 | hCoV-19/Denmark/DCGC-829/2020\|EPI_ISL_614746\|2020- | .................... | .................... | .................... | .................... | .................... | .................... |
| P | 9038 | hCoV-19/Denmark/DCGC-833/2020\|EPI_ISL_614747\|2020- | .................... | .................... | .................... | .................... | .................... | .................... |
| P | 9039 | hCoV-19/Denmark/DCGC-837/2020\|EPI_ISL_614748\|2020- | .................... | .................... | .................... | .................... | .................... | .................... |
| P | 9040 | hCoV-19/Denmark/DCGC-886/2020\|EPI_ISL_614749\|2020- | .................... | .................... | .................... | .................... | .................... | .................... |
| P | 9041 | hCoV-19/Denmark/DCGC-780/2020\|EPI_ISL_614750\|2020- | .................... | .................... | .................... | .................... | .................... | .................... |
| P | 9042 | hCoV-19/Denmark/DCGC-781/2020\|EPI_ISL_614751\|2020- | .................... | .................... | .................... | .................... | .................... | .................... |
| P | 9043 | hCoV-19/Denmark/DCGC-761/2020\|EPI_ISL_614752\|2020- | .................... | .................... | .................... | .................... | .................... | .................... |
| P | 9044 | hCoV-19/Denmark/DCGC-910/2020\|EPI_ISL_614753\|2020- | .................... | .................... | .................... | .................... | .................... | .................... |
| P | 9045 | hCoV-19/Denmark/DCGC-783/2020\|EPI_ISL_614754\|2020- | .................... | .................... | .................... | .................... | .................... | .................... |
| P | 9046 | hCoV-19/Denmark/DCGC-784/2020\|EPI_ISL_614755\|2020- | .................... | .................... | .................... | .................... | .................... | .................... |
| P | 9047 | hCoV-19/Denmark/DCGC-767/2020\|EPI_ISL_614756\|2020- | .................... | .................... | .................... | .................... | .................... | .................... |
| P | 9048 | hCoV-19/Denmark/DCGC-785/2020\|EPI_ISL_614757\|2020- | .................... | .................... | .................... | .................... | .................... | .................... |
| P | 9049 | hCoV-19/Denmark/DCGC-768/2020\|EPI_ISL_614758\|2020- | .................... | .................... | .................... | .................... | .................... | .................... |
| P | 9050 | hCoV-19/Denmark/DCGC-3150/2020\|EPI_ISL_615970\|2020 | .................... | .................... | .................... | .................... | .................... | .................... |
| P | 9051 | hCoV-19/Denmark/DCGC-3151/2020\|EPI_ISL_615971\|2020 | .................... | .................... | .................... | .................... | .................... | .................... |
| P | 9052 | hCoV-19/Denmark/DCGC-3153/2020\|EPI_ISL_615973\|2020 | .................... | .................... | .................... | .................... | .................... | .................... |
| P | 9053 | hCoV-19/Denmark/DCGC-3154/2020\|EPI_ISL_615974\|2020 | .................... | .................... | .................... | .................... | .................... | .................... |
| P | 9054 | hCoV-19/Denmark/DCGC-3155/2020\|EPI_ISL_615975\|2020 | .................... | .................... | .................... | .................... | .................... | .................... |
| P | 9055 | hCoV-19/Denmark/DCGC-3156/2020\|EPI_ISL_615976\|2020 | .................... | .................... | .................... | .................... | .................... | .................... |
| P | 9056 | hCoV-19/Denmark/DCGC-3157/2020\|EPI_ISL_615977\|2020 | .................... | .................... | .................... | .................... | .................... | .................... |
| P | 9057 | hCoV-19/Denmark/DCGC-3158/2020\|EPI_ISL_615978\|2020 | .................... | .................... | .................... | .................... | .................... | .................... |
| P | 9058 | hCoV-19/Denmark/DCGC-3251/2020\|EPI_ISL_615979\|2020 | .................... | .................... | .................... | .................... | .................... | .................... |
| P | 9059 | hCoV-19/Denmark/DCGC-1775/2020\|EPI_ISL_618211\|2020 | .................... | .................... | .................... | .................... | .................... | .................... |
| P | 9060 | hCoV-19/Denmark/DCGC-1789/2020\|EPI_ISL_618220\|2020 | .................... | .................... | .................... | .................... | .................... | .................... |
| P | 9061 | hCoV-19/Denmark/DCGC-1797/2020\|EPI_ISL_618221\|2020 | .................... | .................... | .................... | .................... | .................... | .................... |
| P | 9062 | hCoV-19/Denmark/DCGC-1800/2020\|EPI_ISL_618222\|2020 | .................... | .................... | .................... | .................... | .................... | .................... |
| P | 9063 | hCoV-19/Denmark/DCGC-2014/2020\|EPI_ISL_618224\|2020 | .................... | .................... | .................... | .................... | .................... | .................... |
| P | 9064 | hCoV-19/Denmark/DCGC-2031/2020\|EPI_ISL_618225\|2020 | .................... | .A.................. | .................... | .................... | .................... | .................... |
| P | 9065 | hCoV-19/Denmark/DCGC-2004/2020\|EPI_ISL_618226\|2020 | .................... | .................... | .................... | .................... | .................... | .................... |
| P | 9066 | hCoV-19/Denmark/DCGC-2008/2020\|EPI_ISL_618227\|2020 | .................... | .................... | .................... | .................... | .................... | .................... |
| P | 9067 | hCoV-19/Denmark/DCGC-2012/2020\|EPI_ISL_618228\|2020 | .................... | .................... | .................... | .................... | .................... | .................... |
| P | 9068 | hCoV-19/Denmark/DCGC-2015/2020\|EPI_ISL_618229\|2020 | .................... | .................... | .................... | .................... | .................... | .................... |
| P | 9069 | hCoV-19/Denmark/DCGC-2018/2020\|EPI_ISL_618230\|2020 | .................... | .................... | .................... | .................... | .................... | .................... |
| P | 9070 | hCoV-19/Denmark/DCGC-2021/2020\|EPI_ISL_618231\|2020 | .................... | .................... | .................... | .................... | .................... | .................... |
| P | 9071 | hCoV-19/Denmark/DCGC-1949/2020\|EPI_ISL_618232\|2020 | .................... | .................... | .................... | .................... | .................... | .................... |
| P | 9072 | hCoV-19/Denmark/DCGC-2033/2020\|EPI_ISL_618233\|2020 | .................... | .................... | .................... | .................... | .................... | .................... |
| P | 9073 | hCoV-19/Denmark/DCGC-2002/2020\|EPI_ISL_618234\|2020 | .................... | .................... | .................... | .................... | .................... | .................... |
| P | 9074 | hCoV-19/Denmark/DCGC-2009/2020\|EPI_ISL_618235\|2020 | .................... | .A.................. | .................... | .................... | .................... | .................... |
| P | 9075 | hCoV-19/Denmark/DCGC-2013/2020\|EPI_ISL_618236\|2020 | .................... | .A.................. | .................... | .................... | .................... | .................... |
| P | 9076 | hCoV-19/Denmark/DCGC-2019/2020\|EPI_ISL_618237\|2020 | .................... | .................... | .................... | .................... | .................... | .................... |

| | | | | | | | |
|---|---|---|---|---|---|---|---|
| P | 9077 hCoV-19/Denmark/DCGC-2022/2020\|EPI_ISL_618238\|2020 | ···················· | ···················· | ···················· | ···················· | ···················· | ···················· |
| P | 9078 hCoV-19/Denmark/DCGC-2024/2020\|EPI_ISL_618239\|2020 | ···················· | ···················· | ···················· | ···················· | ···················· | ···················· |
| P | 9079 hCoV-19/Denmark/DCGC-1960/2020\|EPI_ISL_618280\|2020 | ···················· | ···················· | ···················· | ···················· | ···················· | ···················· |
| P | 9080 hCoV-19/Denmark/DCGC-1961/2020\|EPI_ISL_618281\|2020 | ···················· | ···················· | ···················· | ···················· | ···················· | ···················· |
| P | 9081 hCoV-19/Denmark/DCGC-1962/2020\|EPI_ISL_618282\|2020 | ···················· | ···················· | ···················· | ···················· | ···················· | ···················· |
| P | 9082 hCoV-19/Denmark/DCGC-1963/2020\|EPI_ISL_618283\|2020 | ···················· | ···················· | ···················· | ···················· | ···················· | ···················· |
| P | 9083 hCoV-19/Denmark/DCGC-1964/2020\|EPI_ISL_618284\|2020 | ···················· | ···················· | ···················· | ···················· | ···················· | ···················· |
| P | 9084 hCoV-19/Denmark/DCGC-1965/2020\|EPI_ISL_618285\|2020 | ···················· | ···················· | ···················· | ···················· | ···················· | ···················· |
| P | 9085 hCoV-19/Denmark/DCGC-1992/2020\|EPI_ISL_618286\|2020 | ···················· | ···················· | ···················· | ···················· | ···················· | ···················· |
| P | 9086 hCoV-19/Denmark/DCGC-1966/2020\|EPI_ISL_618287\|2020 | ···················· | ·A·················· | ···················· | ···················· | ···················· | ··A················· |
| P | 9087 hCoV-19/Denmark/DCGC-1967/2020\|EPI_ISL_618288\|2020 | ···················· | ···················· | ···················· | ···················· | ···················· | ···················· |
| P | 9088 hCoV-19/Denmark/DCGC-1968/2020\|EPI_ISL_618289\|2020 | ···················· | ···················· | ···················· | ···················· | ···················· | ···················· |
| P | 9089 hCoV-19/Denmark/DCGC-1993/2020\|EPI_ISL_618290\|2020 | ···················· | ···················· | ···················· | ···················· | ···················· | ···················· |
| P | 9090 hCoV-19/Denmark/DCGC-1969/2020\|EPI_ISL_618291\|2020 | ···················· | ···················· | ···················· | ···················· | ···················· | ···················· |
| P | 9091 hCoV-19/Denmark/DCGC-2057/2020\|EPI_ISL_618292\|2020 | ···················· | ···················· | ···················· | ···················· | ···················· | ···················· |
| P | 9092 hCoV-19/Denmark/DCGC-1970/2020\|EPI_ISL_618294\|2020 | ···················· | ···················· | ···················· | ···················· | ···················· | ···················· |
| P | 9093 hCoV-19/Denmark/DCGC-1994/2020\|EPI_ISL_618295\|2020 | ···················· | ···················· | ···················· | ···················· | ···················· | ···················· |
| P | 9094 hCoV-19/Denmark/DCGC-1987/2020\|EPI_ISL_618298\|2020 | ···················· | ···················· | ···················· | ···················· | ···················· | ···················· |
| P | 9095 hCoV-19/Denmark/DCGC-2058/2020\|EPI_ISL_618299\|2020 | ···················· | ···················· | ···················· | ···················· | ···················· | ···················· |
| P | 9096 hCoV-19/Denmark/DCGC-6639/2020\|EPI_ISL_619541\|2020 | ···················· | ···················· | ···················· | ···················· | ···················· | ···················· |
| P | 9097 hCoV-19/Denmark/DCGC-6648/2020\|EPI_ISL_619550\|2020 | ···················· | ···················· | ···················· | ···················· | ···················· | ···················· |
| P | 9098 hCoV-19/Denmark/DCGC-6649/2020\|EPI_ISL_619551\|2020 | ···················· | ···················· | ···················· | ···················· | ···················· | ···················· |
| P | 9099 hCoV-19/Denmark/DCGC-6650/2020\|EPI_ISL_619552\|2020 | ···················· | ···················· | ···················· | ···················· | ···················· | ···················· |
| P | 9100 hCoV-19/Denmark/DCGC-6651/2020\|EPI_ISL_619553\|2020 | ···················· | ···················· | ···················· | ···················· | ···················· | ···················· |
| P | 9101 hCoV-19/Denmark/DCGC-6652/2020\|EPI_ISL_619554\|2020 | ···················· | ···················· | ···················· | ···················· | ···················· | ···················· |
| P | 9102 hCoV-19/Denmark/DCGC-6653/2020\|EPI_ISL_619555\|2020 | ···················· | ···················· | ···················· | ···················· | ···················· | ···················· |
| P | 9103 hCoV-19/Denmark/DCGC-1288/2020\|EPI_ISL_619556\|2020 | ···················· | ···················· | ···················· | ···················· | ···················· | ···················· |
| P | 9104 hCoV-19/Denmark/DCGC-6655/2020\|EPI_ISL_619558\|2020 | ···················· | ···················· | ···················· | ···················· | ···················· | ···················· |
| P | 9105 hCoV-19/Denmark/DCGC-6656/2020\|EPI_ISL_619559\|2020 | ···················· | ·A·················· | ···················· | ···················· | ···················· | ··A················· |
| P | 9106 hCoV-19/Denmark/DCGC-6657/2020\|EPI_ISL_619560\|2020 | ···················· | ···················· | ···················· | ···················· | ···················· | ···················· |
| P | 9107 hCoV-19/Denmark/DCGC-6658/2020\|EPI_ISL_619561\|2020 | ···················· | ···················· | ···················· | ···················· | ···················· | ···················· |
| P | 9108 hCoV-19/Denmark/DCGC-6659/2020\|EPI_ISL_619562\|2020 | ···················· | ···················· | ···················· | ···················· | ···················· | ···················· |
| P | 9109 hCoV-19/Denmark/DCGC-6660/2020\|EPI_ISL_619563\|2020 | ···················· | ···················· | ···················· | ···················· | ···················· | ···················· |
| P | 9110 hCoV-19/Denmark/DCGC-6661/2020\|EPI_ISL_619564\|2020 | ···················· | ···················· | ···················· | ···················· | ···················· | ···················· |
| P | 9111 hCoV-19/Denmark/DCGC-6662/2020\|EPI_ISL_619565\|2020 | ···················· | ···················· | ···················· | ···················· | ···················· | ···················· |
| P | 9112 hCoV-19/Denmark/DCGC-6663/2020\|EPI_ISL_619566\|2020 | ···················· | ···················· | ···················· | ···················· | ···················· | ···················· |
| P | 9113 hCoV-19/Denmark/DCGC-6664/2020\|EPI_ISL_619567\|2020 | ···················· | ···················· | ···················· | ···················· | ···················· | ···················· |
| P | 9114 hCoV-19/Denmark/DCGC-6665/2020\|EPI_ISL_619568\|2020 | ···················· | ···················· | ···················· | ···················· | ···················· | ···················· |
| P | 9115 hCoV-19/Denmark/DCGC-6666/2020\|EPI_ISL_619569\|2020 | ···················· | ···················· | ···················· | ···················· | ···················· | ···················· |
| P | 9116 hCoV-19/Denmark/DCGC-4429/2020\|EPI_ISL_621850\|2020 | ···················· | ···················· | ···················· | ···················· | ···················· | ···················· |
| P | 9117 hCoV-19/Denmark/DCGC-4430/2020\|EPI_ISL_621851\|2020 | ···················· | ···················· | ···················· | ···················· | ···················· | ···················· |
| P | 9118 hCoV-19/Denmark/DCGC-4431/2020\|EPI_ISL_621852\|2020 | ···················· | ···················· | ···················· | ···················· | ···················· | ···················· |
| P | 9119 hCoV-19/Denmark/DCGC-4444/2020\|EPI_ISL_621865\|2020 | ···················· | ···················· | ···················· | ···················· | ···················· | ···················· |
| P | 9120 hCoV-19/Denmark/DCGC-4445/2020\|EPI_ISL_621866\|2020 | ·······T············ | ···················· | ···················· | ···················· | ···················· | ···················· |
| P | 9121 hCoV-19/Denmark/DCGC-4446/2020\|EPI_ISL_621867\|2020 | ···················· | ···················· | ···················· | ···················· | ···················· | ···················· |
| P | 9122 hCoV-19/Denmark/DCGC-4447/2020\|EPI_ISL_621868\|2020 | ·······T············ | ···················· | ···················· | ···················· | ···················· | ···················· |
| P | 9123 hCoV-19/Denmark/DCGC-4448/2020\|EPI_ISL_621869\|2020 | ···················· | ···················· | ···················· | ···················· | ···················· | ···················· |
| P | 9124 hCoV-19/Denmark/DCGC-4449/2020\|EPI_ISL_621870\|2020 | ···················· | ···················· | ···················· | ···················· | ···················· | ···················· |
| P | 9125 hCoV-19/Denmark/DCGC-4450/2020\|EPI_ISL_621871\|2020 | ···················· | ···················· | ···················· | ···················· | ···················· | ···················· |
| P | 9126 hCoV-19/Denmark/DCGC-4451/2020\|EPI_ISL_621872\|2020 | ···················· | ···················· | ···················· | ···················· | ···················· | ···················· |
| P | 9127 hCoV-19/Denmark/DCGC-4452/2020\|EPI_ISL_621873\|2020 | ···················· | ···················· | ···················· | ···················· | ···················· | ···················· |
| P | 9128 hCoV-19/Denmark/DCGC-4642/2020\|EPI_ISL_621874\|2020 | ···················· | ···················· | ···················· | ···················· | ···················· | ···················· |
| P | 9129 hCoV-19/Denmark/DCGC-4643/2020\|EPI_ISL_621875\|2020 | ···················· | ···················· | ···················· | ···················· | ···················· | ···················· |
| P | 9130 hCoV-19/Denmark/DCGC-4644/2020\|EPI_ISL_621876\|2020 | ···················· | ···················· | ···················· | ···················· | ···················· | ···················· |
| P | 9131 hCoV-19/Denmark/DCGC-4645/2020\|EPI_ISL_621877\|2020 | ···················· | ···················· | ···················· | ···················· | ···················· | ···················· |
| P | 9132 hCoV-19/Denmark/DCGC-4646/2020\|EPI_ISL_621878\|2020 | ···················· | ···················· | ···················· | ···················· | ···················· | ···················· |
| P | 9133 hCoV-19/Denmark/DCGC-4647/2020\|EPI_ISL_621879\|2020 | ···················· | ···················· | ···················· | ···················· | ···················· | ···················· |
| P | 9134 hCoV-19/Denmark/DCGC-4648/2020\|EPI_ISL_621880\|2020 | ···················· | ···················· | ···················· | ···················· | ···················· | ···················· |
| P | 9135 hCoV-19/Denmark/DCGC-4650/2020\|EPI_ISL_621882\|2020 | ···················· | ···················· | ···················· | ···················· | ···················· | ···················· |
| P | 9136 hCoV-19/Denmark/DCGC-4652/2020\|EPI_ISL_621884\|2020 | ···················· | ···················· | ···················· | ···················· | ···················· | ···················· |
| P | 9137 hCoV-19/Denmark/DCGC-4653/2020\|EPI_ISL_621885\|2020 | ···················· | ···················· | ···················· | ···················· | ···················· | ···················· |
| P | 9138 hCoV-19/Denmark/DCGC-4654/2020\|EPI_ISL_621886\|2020 | ···················· | ···················· | ···················· | ···················· | ···················· | ···················· |
| P | 9139 hCoV-19/England/ALDP-AA3481/2020\|EPI_ISL_624000\|20 | ···················· | ···················· | ···················· | ···················· | ···················· | ···················· |
| P | 9140 hCoV-19/England/ALDP-AA0F0B/2020\|EPI_ISL_624003\|20 | ···················· | ···················· | ···················· | ···················· | ···················· | ···················· |
| P | 9141 hCoV-19/England/ALDP-AA2DE1/2020\|EPI_ISL_624004\|20 | ···················· | ···················· | ···················· | ···················· | ···················· | ···················· |
| P | 9142 hCoV-19/England/ALDP-AA14CF/2020\|EPI_ISL_624005\|20 | ···················· | ···················· | ···················· | ···················· | ···················· | ···················· |
| P | 9143 hCoV-19/Wales/ALDP-AA24CE/2020\|EPI_ISL_624006\|2020 | ···················· | ···················· | ···················· | ···················· | ···················· | ···················· |
| P | 9144 hCoV-19/England/ALDP-AA0DE3/2020\|EPI_ISL_624008\|20 | ···················· | ···················· | ···················· | ···················· | ···················· | ···················· |
| P | 9145 hCoV-19/England/ALDP-AA51B5/2020\|EPI_ISL_624009\|20 | ···················· | ···················· | ···················· | ···················· | ···················· | ···················· |
| P | 9146 hCoV-19/Scotland/QEUH-AAE574/2020\|EPI_ISL_624010\|2 | ···················· | ···················· | ···················· | ···················· | ···················· | ···················· |
| P | 9147 hCoV-19/England/ALDP-AA13D1/2020\|EPI_ISL_624011\|20 | ···················· | ···················· | ···················· | ···················· | ···················· | ···················· |
| P | 9148 hCoV-19/England/ALDP-AA3779/2020\|EPI_ISL_624013\|20 | ···················· | ···················· | ···················· | ···················· | ···················· | ···················· |
| P | 9149 hCoV-19/England/ALDP-AA075E/2020\|EPI_ISL_624014\|20 | ···················· | ···················· | ···················· | ···················· | ···················· | ···················· |
| P | 9150 hCoV-19/England/ALDP-AA2F81/2020\|EPI_ISL_624015\|20 | ···················· | ·A·················· | ···················· | ···················· | ···················· | ···················· |
| P | 9151 hCoV-19/England/ALDP-AA0F29/2020\|EPI_ISL_624016\|20 | ···················· | ·A·················· | ···················· | ···················· | ···················· | ··A················· |
| P | 9152 hCoV-19/England/ALDP-AA51C4/2020\|EPI_ISL_624017\|20 | ···················· | ···················· | ···················· | ···················· | ···················· | ···················· |
| P | 9153 hCoV-19/Scotland/QEUH-AAE8F3/2020\|EPI_ISL_624018\|2 | ···················· | ···················· | ···················· | ···················· | ···················· | ···················· |
| P | 9154 hCoV-19/England/QEUH-AAE42C/2020\|EPI_ISL_624019\|20 | ···················· | ···················· | ···················· | ···················· | ···················· | ···················· |
| P | 9155 hCoV-19/England/ALDP-AA0703/2020\|EPI_ISL_624020\|20 | ···················· | ···················· | ···················· | ···················· | ···················· | ···················· |
| P | 9156 hCoV-19/England/ALDP-AA416B/2020\|EPI_ISL_624021\|20 | ···················· | ···················· | ···················· | ···················· | ···················· | ···················· |

| | | | | | | | |
|---|---|---|---|---|---|---|---|
| P | 9157 hCoV-19/Scotland/QEUH-AADD8C/2020\|EPI_ISL_624022\|2 | ···················· | ···················· | ···················· | ···················· | ···················· | ···················· |
| P | 9158 hCoV-19/England/ALDP-AA0F47/2020\|EPI_ISL_624023\|20 | ···················· | ·A·················· | ···················· | ···················· | ···················· | ···A················ |
| P | 9159 hCoV-19/England/ALDP-AA3490/2020\|EPI_ISL_624025\|20 | ···················· | ···················· | ···················· | ···················· | ···················· | ···················· |
| P | 9160 hCoV-19/England/ALDP-AA134A/2020\|EPI_ISL_624026\|20 | ···················· | ···················· | ···················· | ···················· | ···················· | ···················· |
| P | 9161 hCoV-19/England/QEUH-AAD6FA/2020\|EPI_ISL_624027\|20 | ···················· | ···················· | ···················· | ···················· | ···················· | ···················· |
| P | 9162 hCoV-19/Scotland/QEUH-AAD35D/2020\|EPI_ISL_624028\|2 | ···················· | ···················· | ···················· | ···················· | ···················· | ···················· |
| P | 9163 hCoV-19/England/ALDP-AA2B23/2020\|EPI_ISL_624030\|20 | ···················· | ···················· | ···················· | ···················· | ···················· | ···················· |
| P | 9164 hCoV-19/England/ALDP-AA2E39/2020\|EPI_ISL_624031\|20 | ···················· | ···················· | ···················· | ···················· | ···················· | ···················· |
| P | 9165 hCoV-19/Scotland/QEUH-AAE4E0/2020\|EPI_ISL_624032\|2 | ···················· | ···················· | ···················· | ···················· | ···················· | ···················· |
| P | 9166 hCoV-19/Scotland/QEUH-AAEEB5/2020\|EPI_ISL_624034\|2 | ···················· | ···················· | ···················· | ···················· | ···················· | ···············A···· |
| P | 9167 hCoV-19/England/ALDP-AA1632/2020\|EPI_ISL_624035\|20 | ···················· | ···················· | ···················· | ···················· | ···················· | ···················· |
| P | 9168 hCoV-19/England/ALDP-AA2F18/2020\|EPI_ISL_624036\|20 | ···················· | ···················· | ···················· | ···················· | ···················· | ···················· |
| P | 9169 hCoV-19/England/ALDP-AA4110/2020\|EPI_ISL_624037\|20 | ···················· | ···················· | ···················· | ···················· | ···················· | ···················· |
| P | 9170 hCoV-19/England/ALDP-AA19A2/2020\|EPI_ISL_624038\|20 | ···················· | ···················· | ···················· | ···················· | ···················· | ···················· |
| P | 9171 hCoV-19/England/ALDP-AA414D/2020\|EPI_ISL_624039\|20 | ···················· | ···················· | ···················· | ···················· | ···················· | ···················· |
| P | 9172 hCoV-19/Wales/CAMC-A82066/2020\|EPI_ISL_625300\|2020 | ···················· | ···················· | ···················· | ···················· | ···················· | ···················· |
| P | 9173 hCoV-19/Wales/CAMC-A8245B/2020\|EPI_ISL_625301\|2020 | ···················· | ···················· | ···················· | ···················· | ···················· | ···················· |
| P | 9174 hCoV-19/Scotland/CAMC-A82C44/2020\|EPI_ISL_625302\|2 | ···················· | ···················· | ···················· | ···················· | ···················· | ···················· |
| P | 9175 hCoV-19/Wales/CAMC-A82321/2020\|EPI_ISL_625303\|2020 | ···················· | ···················· | ···················· | ···················· | ···················· | ···················· |
| P | 9176 hCoV-19/Wales/CAMC-A80E22/2020\|EPI_ISL_625305\|2020 | ···················· | ···················· | ···················· | ···················· | ···················· | ···················· |
| P | 9177 hCoV-19/Wales/CAMC-A81E5E/2020\|EPI_ISL_625306\|2020 | ···················· | ···················· | ···················· | ···················· | ···················· | ···················· |
| P | 9178 hCoV-19/Scotland/CAMC-A8236D/2020\|EPI_ISL_625307\|2 | ···················· | ···················· | ···················· | ···················· | ···················· | ···················· |
| P | 9179 hCoV-19/Scotland/QEUH-A9EDAC/2020\|EPI_ISL_625330\|2 | ···················· | ···················· | ···················· | ···················· | ···················· | ···················· |
| P | 9180 hCoV-19/Scotland/QEUH-A9E9F3/2020\|EPI_ISL_625332\|2 | ···················· | ···················· | ···················· | ···················· | ···················· | ···················· |
| P | 9181 hCoV-19/Scotland/QEUH-A9E823/2020\|EPI_ISL_625333\|2 | ···················· | ···················· | ···················· | ···················· | ···················· | ···················· |
| P | 9182 hCoV-19/Scotland/QEUH-A9B404/2020\|EPI_ISL_625335\|2 | ···················· | ···················· | ···················· | ···················· | ···················· | ···················· |
| P | 9183 hCoV-19/Scotland/QEUH-A9E920/2020\|EPI_ISL_625336\|2 | ···················· | ···················· | ···················· | ···················· | ···················· | ···················· |
| P | 9184 hCoV-19/Scotland/QEUH-A9E87E/2020\|EPI_ISL_625337\|2 | ···················· | ···················· | ···················· | ···················· | ···················· | ···················· |
| P | 9185 hCoV-19/Scotland/QEUH-A9E832/2020\|EPI_ISL_625338\|2 | ···················· | ······A············· | ···················· | ···················· | ···················· | ···················· |
| P | 9186 hCoV-19/Scotland/QEUH-A9B45F/2020\|EPI_ISL_625339\|2 | ···················· | ···················· | ···················· | ···················· | ···················· | ···················· |
| P | 9187 hCoV-19/Scotland/QEUH-A9B905/2020\|EPI_ISL_625340\|2 | ···················· | ···················· | ···················· | ···················· | ···················· | ···················· |
| P | 9188 hCoV-19/Wales/QEUH-A9FB92/2020\|EPI_ISL_625341\|2020 | ···················· | ···················· | ···················· | ···················· | ···················· | ···················· |
| P | 9189 hCoV-19/Scotland/QEUH-A9EA5A/2020\|EPI_ISL_625342\|2 | ···················· | ···················· | ···················· | ···················· | ···················· | ···················· |
| P | 9190 hCoV-19/Scotland/QEUH-A9F084/2020\|EPI_ISL_625343\|2 | ···················· | ···················· | ···················· | ···················· | ···················· | ···················· |
| P | 9191 hCoV-19/Scotland/QEUH-A9EAC3/2020\|EPI_ISL_625344\|2 | ···················· | ···················· | ···················· | ···················· | ···················· | ···················· |
| P | 9192 hCoV-19/Scotland/QEUH-A9B3CB/2020\|EPI_ISL_625345\|2 | ···················· | ···················· | ···················· | ···················· | ···················· | ···················· |
| P | 9193 hCoV-19/Scotland/QEUH-A9EA69/2020\|EPI_ISL_625346\|2 | ···················· | ···················· | ···················· | ···················· | ···················· | ···················· |
| P | 9194 hCoV-19/Scotland/QEUH-A9B370/2020\|EPI_ISL_625347\|2 | ···················· | ···················· | ···················· | ···················· | ···················· | ···················· |
| P | 9195 hCoV-19/Scotland/QEUH-A9E96C/2020\|EPI_ISL_625348\|2 | ···················· | ···················· | ···················· | ···················· | ···················· | ···················· |
| P | 9196 hCoV-19/Scotland/QEUH-A9E86F/2020\|EPI_ISL_625349\|2 | ···················· | ···················· | ···················· | ···················· | ···················· | ···················· |
| P | 9197 hCoV-19/Scotland/QEUH-A9B422/2020\|EPI_ISL_625350\|2 | ···················· | ···················· | ···················· | ···················· | ···················· | ···················· |
| P | 9198 hCoV-19/Scotland/QEUH-A9E94E/2020\|EPI_ISL_625351\|2 | ···················· | ···················· | ···················· | ···················· | ···················· | ···················· |
| P | 9199 hCoV-19/Scotland/QEUH-A9B64A/2020\|EPI_ISL_625352\|2 | ···················· | ···················· | ···················· | ···················· | ···················· | ···················· |
| P | 9200 hCoV-19/Scotland/QEUH-A9E8E7/2020\|EPI_ISL_625353\|2 | ···················· | ···················· | ···················· | ···················· | ···················· | ···················· |
| P | 9201 hCoV-19/Scotland/QEUH-A9B343/2020\|EPI_ISL_625354\|2 | ···················· | ···················· | ···················· | ···················· | ···················· | ···················· |
| P | 9202 hCoV-19/Scotland/QEUH-A9EA1E/2020\|EPI_ISL_625355\|2 | ···················· | ···················· | ···················· | ···················· | ···················· | ···················· |
| P | 9203 hCoV-19/Scotland/QEUH-A9EAB4/2020\|EPI_ISL_625356\|2 | ···················· | ···················· | ···················· | ···················· | ···················· | ···················· |
| P | 9204 hCoV-19/Scotland/QEUH-A9B63B/2020\|EPI_ISL_625357\|2 | ···················· | ···················· | ···················· | ···················· | ···················· | ···················· |
| P | 9205 hCoV-19/Scotland/QEUH-A9EFF1/2020\|EPI_ISL_625359\|2 | ···················· | ···················· | ···················· | ···················· | ···················· | ···················· |
| P | 9206 hCoV-19/Scotland/QEUH-A9B46E/2020\|EPI_ISL_625361\|2 | ···················· | ···················· | ···················· | ···················· | ···················· | ···················· |
| P | 9207 hCoV-19/Scotland/QEUH-A9E95D/2020\|EPI_ISL_625362\|2 | ···················· | ···················· | ···················· | ···················· | ···················· | ···················· |
| P | 9208 hCoV-19/Scotland/QEUH-A9B3F8/2020\|EPI_ISL_625363\|2 | ···················· | ···················· | ···················· | ···················· | ···················· | ···················· |
| P | 9209 hCoV-19/Scotland/QEUH-A9AE34/2020\|EPI_ISL_625364\|2 | ···················· | ···················· | ···················· | ···················· | ···················· | ···················· |
| P | 9210 hCoV-19/Scotland/QEUH-A9B49B/2020\|EPI_ISL_625366\|2 | ···················· | ···················· | ···················· | ···················· | ···················· | ···················· |
| P | 9211 hCoV-19/Scotland/QEUH-A9B334/2020\|EPI_ISL_625367\|2 | ···················· | ···················· | ···················· | ···················· | ···················· | ···················· |
| P | 9212 hCoV-19/Scotland/QEUH-A9EDF7/2020\|EPI_ISL_625368\|2 | ···················· | ···················· | ···················· | ···················· | ···················· | ···················· |
| P | 9213 hCoV-19/Scotland/QEUH-A9EAA5/2020\|EPI_ISL_625369\|2 | ···················· | ···················· | ···················· | ···················· | ···················· | ···················· |
| P | 9214 hCoV-19/Wales/ALDP-A8635A/2020\|EPI_ISL_625400\|2020 | ···················· | ···················· | ···················· | ···················· | ···················· | ···················· |
| P | 9215 hCoV-19/Wales/ALDP-A700E3/2020\|EPI_ISL_625403\|2020 | ···················· | ···················· | ···················· | ···················· | ···················· | ···················· |
| P | 9216 hCoV-19/Scotland/MILK-A6A046/2020\|EPI_ISL_625406\|2 | ···················· | ···················· | ···················· | ···················· | ···················· | ···················· |
| P | 9217 hCoV-19/Denmark/DCGC-1166/2020\|EPI_ISL_614638\|2020 | ···················· | ···················· | ···················· | ···················· | ···················· | ···················· |
| P | 9218 hCoV-19/Denmark/DCGC-1143/2020\|EPI_ISL_614649\|2020 | ···················· | ···················· | ···················· | ···················· | ···················· | ···················· |
| P | 9219 hCoV-19/Denmark/DCGC-1153/2020\|EPI_ISL_614650\|2020 | ···················· | ···················· | ···················· | ···················· | ···················· | ···················· |
| P | 9220 hCoV-19/Denmark/DCGC-1157/2020\|EPI_ISL_614651\|2020 | ···················· | ···················· | ···················· | ···················· | ···················· | ···················· |
| P | 9221 hCoV-19/Denmark/DCGC-1163/2020\|EPI_ISL_614652\|2020 | ···················· | ···················· | ···················· | ···················· | ···················· | ···················· |
| P | 9222 hCoV-19/Denmark/DCGC-1167/2020\|EPI_ISL_614653\|2020 | ···················· | ···················· | ···················· | ···················· | ···················· | ···················· |
| P | 9223 hCoV-19/Denmark/DCGC-1173/2020\|EPI_ISL_614654\|2020 | ···················· | ···················· | ···················· | ···················· | ···················· | ···················· |
| P | 9224 hCoV-19/Denmark/DCGC-1179/2020\|EPI_ISL_614655\|2020 | ···················· | ···················· | ···················· | ···················· | ···················· | ···················· |
| P | 9225 hCoV-19/Denmark/DCGC-1140/2020\|EPI_ISL_614656\|2020 | ···················· | ···················· | ···················· | ···················· | ···················· | ···················· |
| P | 9226 hCoV-19/Denmark/DCGC-1144/2020\|EPI_ISL_614657\|2020 | ···················· | ···················· | ···················· | ···················· | ···················· | ···················· |
| P | 9227 hCoV-19/Denmark/DCGC-1148/2020\|EPI_ISL_614658\|2020 | ···················· | ···················· | ···················· | ···················· | ···················· | ···················· |
| P | 9228 hCoV-19/Denmark/DCGC-1154/2020\|EPI_ISL_614659\|2020 | ···················· | ···················· | ···················· | ···················· | ···················· | ···················· |
| P | 9229 hCoV-19/Denmark/DCGC-1158/2020\|EPI_ISL_614660\|2020 | ···················· | ···················· | ···················· | ···················· | ···················· | ···················· |
| P | 9230 hCoV-19/Denmark/DCGC-1164/2020\|EPI_ISL_614661\|2020 | ···················· | ···················· | ···················· | ···················· | ···················· | ···················· |
| P | 9231 hCoV-19/Denmark/DCGC-1168/2020\|EPI_ISL_614662\|2020 | ···················· | ···················· | ···················· | ···················· | ···················· | ···················· |
| P | 9232 hCoV-19/Denmark/DCGC-928/2020\|EPI_ISL_614663\|2020- | ···················· | ···················· | ···················· | ···················· | ···················· | ···················· |
| P | 9233 hCoV-19/Denmark/DCGC-1170/2020\|EPI_ISL_614664\|2020 | ···················· | ···················· | ···················· | ···················· | ···················· | ···················· |
| P | 9234 hCoV-19/Denmark/DCGC-1174/2020\|EPI_ISL_614665\|2020 | ···················· | ···················· | ···················· | ···················· | ···················· | ···················· |
| P | 9235 hCoV-19/Denmark/DCGC-1180/2020\|EPI_ISL_614666\|2020 | ···················· | ···················· | ···················· | ···················· | ···················· | ···················· |
| P | 9236 hCoV-19/Denmark/DCGC-1149/2020\|EPI_ISL_614667\|2020 | ···················· | ···················· | ···················· | ···················· | ···················· | ···················· |

| | | | | | | | | |
|---|---|---|---|---|---|---|---|---|
| P | 9237 | hCoV-19/Denmark/DCGC-1159/2020\|EPI_ISL_614668\|2020 | .................... | .................... | ..................... | .................... | .................... | .................... |
| P | 9238 | hCoV-19/Denmark/DCGC-931/2020\|EPI_ISL_614669\|2020- | .................... | .................... | ..................... | .................... | .................... | .................... |
| P | 9239 | hCoV-19/Denmark/DCGC-1175/2020\|EPI_ISL_614670\|2020 | .................... | .................... | ..................... | .................... | .................... | .................... |
| P | 9240 | hCoV-19/Denmark/DCGC-1039/2020\|EPI_ISL_614671\|2020 | .................... | .................... | ..................... | .................... | .................... | .................... |
| P | 9241 | hCoV-19/Denmark/DCGC-1040/2020\|EPI_ISL_614672\|2020 | .................... | .................... | ..................... | .................... | .................... | .................... |
| P | 9242 | hCoV-19/Denmark/DCGC-1041/2020\|EPI_ISL_614673\|2020 | .................... | .................... | ..................... | .................... | .................... | .................... |
| P | 9243 | hCoV-19/Denmark/DCGC-1043/2020\|EPI_ISL_614674\|2020 | .................... | .................... | ..................... | .................... | .................... | .................... |
| P | 9244 | hCoV-19/Denmark/DCGC-1044/2020\|EPI_ISL_614675\|2020 | .................... | .................... | ..................... | .................... | .................... | .................... |
| P | 9245 | hCoV-19/Denmark/DCGC-1045/2020\|EPI_ISL_614676\|2020 | .................... | .................... | ..................... | .................... | .................... | .................... |
| P | 9246 | hCoV-19/Denmark/DCGC-1046/2020\|EPI_ISL_614677\|2020 | .................... | .................... | ..................... | .................... | .................... | .................... |
| P | 9247 | hCoV-19/Denmark/DCGC-1047/2020\|EPI_ISL_614678\|2020 | .................... | .................... | ..................... | .................... | .................... | .................... |
| P | 9248 | hCoV-19/Denmark/DCGC-1048/2020\|EPI_ISL_614679\|2020 | .................... | .................... | ..................... | .................... | .................... | .................... |
| P | 9249 | hCoV-19/Denmark/DCGC-3149/2020\|EPI_ISL_615969\|2020 | .................... | .................... | ..................... | .................... | .................... | .................... |
| P | 9250 | hCoV-19/Denmark/DCGC-3160/2020\|EPI_ISL_615981\|2020 | .................... | .................... | ..................... | .................... | .................... | .................... |
| P | 9251 | hCoV-19/Denmark/DCGC-3252/2020\|EPI_ISL_615982\|2020 | .................... | .................... | ..................... | .................... | .................... | .................... |
| P | 9252 | hCoV-19/Denmark/DCGC-3162/2020\|EPI_ISL_615984\|2020 | .................... | .................... | ..................... | .................... | .................... | .................... |
| P | 9253 | hCoV-19/Denmark/DCGC-3163/2020\|EPI_ISL_615985\|2020 | .................... | .................... | ..................... | .................... | .................... | .................... |
| P | 9254 | hCoV-19/Denmark/DCGC-3164/2020\|EPI_ISL_615986\|2020 | .................... | .................... | ..................... | .................... | .................... | .................... |
| P | 9255 | hCoV-19/Denmark/DCGC-3165/2020\|EPI_ISL_615987\|2020 | .................... | .................... | ..................... | .................... | .................... | .................... |
| P | 9256 | hCoV-19/Denmark/DCGC-3167/2020\|EPI_ISL_615989\|2020 | .................... | .................... | ..................... | .................... | .................... | .................... |
| P | 9257 | hCoV-19/Denmark/DCGC-3169/2020\|EPI_ISL_615991\|2020 | .................... | .................... | ..................... | .................... | .................... | .................... |
| P | 9258 | hCoV-19/Denmark/DCGC-3170/2020\|EPI_ISL_615992\|2020 | .................... | .................... | ..................... | .................... | .................... | .................... |
| P | 9259 | hCoV-19/Denmark/DCGC-3171/2020\|EPI_ISL_615993\|2020 | .................... | .................... | ..................... | .................... | .................... | .................... |
| P | 9260 | hCoV-19/Denmark/DCGC-3172/2020\|EPI_ISL_615994\|2020 | .................... | .................... | ..................... | .................... | .................... | .................... |
| P | 9261 | hCoV-19/Denmark/DCGC-3173/2020\|EPI_ISL_615995\|2020 | .................... | .................... | ..................... | .................... | .................... | .................... |
| P | 9262 | hCoV-19/Denmark/DCGC-3175/2020\|EPI_ISL_615997\|2020 | .................... | .................... | ..................... | .................... | .................... | .................... |
| P | 9263 | hCoV-19/Denmark/DCGC-3176/2020\|EPI_ISL_615998\|2020 | .................... | .................... | ..................... | .................... | .................... | .................... |
| P | 9264 | hCoV-19/Denmark/DCGC-3177/2020\|EPI_ISL_615999\|2020 | .................... | .................... | ..................... | .................... | .................... | .................... |
| P | 9265 | hCoV-19/Denmark/DCGC-1516/2020\|EPI_ISL_618120\|2020 | .................... | .................... | ..................... | .................... | .................... | .................... |
| P | 9266 | hCoV-19/Denmark/DCGC-1517/2020\|EPI_ISL_618121\|2020 | .................... | .................... | ..................... | .................... | .................... | .................... |
| P | 9267 | hCoV-19/Denmark/DCGC-1518/2020\|EPI_ISL_618122\|2020 | .................... | .................... | ..................... | .................... | .................... | .................... |
| P | 9268 | hCoV-19/Denmark/DCGC-1520/2020\|EPI_ISL_618124\|2020 | .................... | .................... | ..................... | .................... | .................... | .................... |
| P | 9269 | hCoV-19/Denmark/DCGC-1521/2020\|EPI_ISL_618125\|2020 | .................... | .................... | ..................... | .................... | .................... | .................... |
| P | 9270 | hCoV-19/Denmark/DCGC-1524/2020\|EPI_ISL_618128\|2020 | .................... | .................... | ..................... | .................... | .................... | .................... |
| P | 9271 | hCoV-19/Denmark/DCGC-1525/2020\|EPI_ISL_618129\|2020 | .................... | .................... | ..................... | .................... | .................... | .................... |
| P | 9272 | hCoV-19/Denmark/DCGC-1526/2020\|EPI_ISL_618130\|2020 | .................... | .................... | ..................... | .................... | .................... | .................... |
| P | 9273 | hCoV-19/Denmark/DCGC-1528/2020\|EPI_ISL_618132\|2020 | .................... | .................... | ..................... | .................... | .................... | .................... |
| P | 9274 | hCoV-19/Denmark/DCGC-1529/2020\|EPI_ISL_618133\|2020 | .................... | .................... | ..................... | .................... | .................... | .................... |
| P | 9275 | hCoV-19/Denmark/DCGC-1530/2020\|EPI_ISL_618134\|2020 | .................... | .................... | ..................... | .................... | .................... | .................... |
| P | 9276 | hCoV-19/Denmark/DCGC-1531/2020\|EPI_ISL_618135\|2020 | .................... | .................... | ..................... | .................... | .................... | .................... |
| P | 9277 | hCoV-19/Denmark/DCGC-1532/2020\|EPI_ISL_618136\|2020 | .................... | .................... | ..................... | .................... | .................... | .................... |
| P | 9278 | hCoV-19/Denmark/DCGC-1567/2020\|EPI_ISL_618137\|2020 | .................... | .................... | ..................... | .................... | .................... | .................... |
| P | 9279 | hCoV-19/Denmark/DCGC-1569/2020\|EPI_ISL_618139\|2020 | .................... | .................... | ..................... | .................... | .................... | .................... |
| P | 9280 | hCoV-19/Denmark/DCGC-1576/2020\|EPI_ISL_618146\|2020 | .................... | .................... | ..................... | .................... | .................... | .................... |
| P | 9281 | hCoV-19/Denmark/DCGC-1577/2020\|EPI_ISL_618147\|2020 | .................... | .................... | ..................... | .................... | .................... | .................... |
| P | 9282 | hCoV-19/Denmark/DCGC-6550/2020\|EPI_ISL_619450\|2020 | .................... | .................... | ..................... | .................... | .................... | .................... |
| P | 9283 | hCoV-19/Denmark/DCGC-6551/2020\|EPI_ISL_619451\|2020 | .................... | .................... | ..................... | .................... | .................... | .................... |
| P | 9284 | hCoV-19/Denmark/DCGC-6552/2020\|EPI_ISL_619452\|2020 | .................... | .................... | ..................... | .................... | .................... | .................... |
| P | 9285 | hCoV-19/Denmark/DCGC-6553/2020\|EPI_ISL_619453\|2020 | .................... | .................... | ..................... | .................... | .................... | .................... |
| P | 9286 | hCoV-19/Denmark/DCGC-6554/2020\|EPI_ISL_619454\|2020 | .................... | .................... | ..................... | .................... | .................... | .................... |
| P | 9287 | hCoV-19/Denmark/DCGC-1285/2020\|EPI_ISL_619455\|2020 | .................... | .................... | ..................... | .................... | .................... | .................... |
| P | 9288 | hCoV-19/Denmark/DCGC-1809/2020\|EPI_ISL_619456\|2020 | .................... | .................... | ..................... | .................... | .................... | .................... |
| P | 9289 | hCoV-19/Denmark/DCGC-6763/2020\|EPI_ISL_619459\|2020 | .................... | .................... | ..................... | .................... | .................... | .................... |
| P | 9290 | hCoV-19/Denmark/DCGC-6765/2020\|EPI_ISL_619461\|2020 | .................... | .................... | ..................... | .................... | .................... | .................... |
| P | 9291 | hCoV-19/Denmark/DCGC-6766/2020\|EPI_ISL_619462\|2020 | .................... | .................... | ..................... | .................... | .................... | .................... |
| P | 9292 | hCoV-19/Denmark/DCGC-6769/2020\|EPI_ISL_619465\|2020 | .................... | .................... | ..................... | .................... | .................... | .................... |
| P | 9293 | hCoV-19/Denmark/DCGC-6770/2020\|EPI_ISL_619466\|2020 | .................... | .................... | ..................... | .................... | .................... | .................... |
| P | 9294 | hCoV-19/Denmark/DCGC-6772/2020\|EPI_ISL_619468\|2020 | .................... | .................... | ..................... | .................... | .................... | .................... |
| P | 9295 | hCoV-19/Denmark/DCGC-6781/2020\|EPI_ISL_619477\|2020 | .................... | .................... | ..................... | .................... | .................... | .................... |
| P | 9296 | hCoV-19/Denmark/DCGC-6782/2020\|EPI_ISL_619478\|2020 | .................... | .................... | ..................... | .................... | .................... | .................... |
| P | 9297 | hCoV-19/Denmark/DCGC-6803/2020\|EPI_ISL_619500\|2020 | .................... | .................... | ..................... | .................... | .................... | .................... |
| P | 9298 | hCoV-19/Denmark/DCGC-6804/2020\|EPI_ISL_619501\|2020 | .................... | .................... | ..................... | .................... | .................... | .................... |
| P | 9299 | hCoV-19/Denmark/DCGC-6805/2020\|EPI_ISL_619502\|2020 | .................... | .................... | ..................... | .................... | .................... | .................... |
| P | 9300 | hCoV-19/Denmark/DCGC-6806/2020\|EPI_ISL_619503\|2020 | .................... | .................... | ..................... | .................... | .................... | .................... |
| P | 9301 | hCoV-19/Denmark/DCGC-6807/2020\|EPI_ISL_619504\|2020 | .................... | .................... | ..................... | .................... | .................... | .................... |
| P | 9302 | hCoV-19/Denmark/DCGC-6808/2020\|EPI_ISL_619505\|2020 | .................... | .................... | ..................... | .................... | .................... | .................... |
| P | 9303 | hCoV-19/Denmark/DCGC-6811/2020\|EPI_ISL_619508\|2020 | .................... | .................... | ..................... | .................... | .................... | .................... |
| P | 9304 | hCoV-19/Denmark/DCGC-6812/2020\|EPI_ISL_619509\|2020 | .................... | .................... | ..................... | .................... | .................... | .................... |
| P | 9305 | hCoV-19/Denmark/DCGC-6813/2020\|EPI_ISL_619510\|2020 | .................... | .................... | ..................... | .................... | .................... | .................... |
| P | 9306 | hCoV-19/Denmark/DCGC-6816/2020\|EPI_ISL_619513\|2020 | .................... | .................... | ..................... | .................... | .................... | .................... |
| P | 9307 | hCoV-19/Denmark/DCGC-6818/2020\|EPI_ISL_619515\|2020 | .................... | .................... | ..................... | .................... | .................... | .................... |
| P | 9308 | hCoV-19/Denmark/DCGC-6820/2020\|EPI_ISL_619517\|2020 | .................... | .................... | ..................... | .................... | .................... | .................... |
| P | 9309 | hCoV-19/Denmark/DCGC-6822/2020\|EPI_ISL_619519\|2020 | .................... | .................... | ..................... | .................... | .................... | .................... |
| P | 9310 | hCoV-19/Denmark/DCGC-6823/2020\|EPI_ISL_619520\|2020 | .................... | .................... | ..................... | .................... | .................... | .................... |
| P | 9311 | hCoV-19/Denmark/DCGC-6824/2020\|EPI_ISL_619521\|2020 | .................... | .................... | ..................... | .................... | .................... | .................... |
| P | 9312 | hCoV-19/Denmark/DCGC-6825/2020\|EPI_ISL_619522\|2020 | .................... | .................... | ..................... | .................... | .................... | .................... |
| P | 9313 | hCoV-19/Denmark/DCGC-6826/2020\|EPI_ISL_619523\|2020 | .................... | .................... | ..................... | .................... | .................... | .................... |
| P | 9314 | hCoV-19/Denmark/DCGC-6827/2020\|EPI_ISL_619524\|2020 | .................... | .................... | ..................... | .................... | .................... | .................... |
| P | 9315 | hCoV-19/Denmark/DCGC-6829/2020\|EPI_ISL_619526\|2020 | .................... | .................... | ..................... | .................... | .................... | .................... |
| P | 9316 | hCoV-19/Denmark/DCGC-6831/2020\|EPI_ISL_619528\|2020 | .................... | .................... | ..................... | .................... | .................... | .................... |

```
P 9317 hCoV-19/Denmark/DCGC-6832/2020|EPI_ISL_619529|2020 .................... .................... .................... .................... .................... ....................
P 9318 hCoV-19/Denmark/DCGC-5040/2020|EPI_ISL_621766|2020 .................... .................... .................... .................... .................... ....................
P 9319 hCoV-19/Denmark/DCGC-5047/2020|EPI_ISL_621767|2020 .................... .................... .................... .................... .................... ....................
P 9320 hCoV-19/Denmark/DCGC-4979/2020|EPI_ISL_621768|2020 .................... .................... .................... .................... .................... ....................
P 9321 hCoV-19/Denmark/DCGC-4985/2020|EPI_ISL_621769|2020 .................... .................... .................... .................... .................... ....................
P 9322 hCoV-19/Denmark/DCGC-4993/2020|EPI_ISL_621770|2020 .................... .................... .................... .................... .................... ....................
P 9323 hCoV-19/Denmark/DCGC-5004/2020|EPI_ISL_621771|2020 .................... .................... .................... .................... .................... ....................
P 9324 hCoV-19/Denmark/DCGC-5010/2020|EPI_ISL_621772|2020 .................... .................... .................... .................... .................... ....................
P 9325 hCoV-19/Denmark/DCGC-5016/2020|EPI_ISL_621773|2020 .................... .................... .................... .................... .................... ....................
P 9326 hCoV-19/Denmark/DCGC-5023/2020|EPI_ISL_621774|2020 .................... .................... .................... .................... .................... ....................
P 9327 hCoV-19/Denmark/DCGC-5041/2020|EPI_ISL_621775|2020 .................... .................... .................... .................... .................... ....................
P 9328 hCoV-19/Denmark/DCGC-5048/2020|EPI_ISL_621776|2020 .................... .................... .................... .................... .................... ....................
P 9329 hCoV-19/Denmark/DCGC-4980/2020|EPI_ISL_621777|2020 .................... .................... .................... .................... ......T............. ....................
P 9330 hCoV-19/Denmark/DCGC-1653/2020|EPI_ISL_621778|2020 .................... .................... .................... .................... .................... ....................
P 9331 hCoV-19/Denmark/DCGC-4986/2020|EPI_ISL_621779|2020 .................... .................... .................... .................... .................... ....................
P 9332 hCoV-19/Denmark/DCGC-4994/2020|EPI_ISL_621780|2020 .................... .................... .................... .................... .................... ....................
P 9333 hCoV-19/Denmark/DCGC-4999/2020|EPI_ISL_621781|2020 .................... .................... .................... .................... .................... ....................
P 9334 hCoV-19/Denmark/DCGC-5005/2020|EPI_ISL_621782|2020 .................... .................... .................... .................... .................... ....................
P 9335 hCoV-19/Denmark/DCGC-5017/2020|EPI_ISL_621783|2020 .................... .................... .................... .................... .................... ....................
P 9336 hCoV-19/Denmark/DCGC-5024/2020|EPI_ISL_621784|2020 .................... .................... .................... .................... .................... ....................
P 9337 hCoV-19/Denmark/DCGC-5030/2020|EPI_ISL_621785|2020 .................... .................... .................... .................... .................... ....................
P 9338 hCoV-19/Denmark/DCGC-5042/2020|EPI_ISL_621786|2020 .................... .................... .................... .................... .................... ....................
P 9339 hCoV-19/Denmark/DCGC-4981/2020|EPI_ISL_621787|2020 .................... .................... .................... .................... .................... ....................
P 9340 hCoV-19/Denmark/DCGC-4996/2020|EPI_ISL_621799|2020 .................... .................... .................... .................... .................... ....................
P 9341 hCoV-19/Denmark/DCGC-5007/2020|EPI_ISL_621800|2020 .................... .................... .................... .................... .................... ....................
P 9342 hCoV-19/Denmark/DCGC-5012/2020|EPI_ISL_621801|2020 .................... .................... .................... .................... .................... ....................
P 9343 hCoV-19/Denmark/DCGC-5019/2020|EPI_ISL_621802|2020 .................... .................... .................... .................... .................... ....................
P 9344 hCoV-19/Denmark/DCGC-5026/2020|EPI_ISL_621803|2020 .................... .................... .................... .................... .................... ....................
P 9345 hCoV-19/Denmark/DCGC-5037/2020|EPI_ISL_621804|2020 .................... .................... .................... .................... .................... ....................
P 9346 hCoV-19/Denmark/DCGC-4982/2020|EPI_ISL_621805|2020 .................... .................... .................... .................... .................... ....................
P 9347 hCoV-19/Denmark/DCGC-4989/2020|EPI_ISL_621806|2020 .................... .................... .................... .................... .................... ....................
P 9348 hCoV-19/Denmark/DCGC-4997/2020|EPI_ISL_621807|2020 .................... .................... .................... .................... .................... ....................
P 9349 hCoV-19/Denmark/DCGC-5000/2020|EPI_ISL_621808|2020 .................... .................... .................... .................... .................... ....................
P 9350 hCoV-19/Denmark/DCGC-5013/2020|EPI_ISL_621809|2020 .................... .................... .................... .................... .................... ....................
P 9351 hCoV-19/Denmark/DCGC-5020/2020|EPI_ISL_621810|2020 .................... .................... .................... .................... .................... ....................
P 9352 hCoV-19/Denmark/DCGC-5032/2020|EPI_ISL_621811|2020 .................... .................... .................... .................... .................... ....................
P 9353 hCoV-19/Denmark/DCGC-5038/2020|EPI_ISL_621812|2020 .................... .................... .................... .................... .................... ....................
P 9354 hCoV-19/Denmark/DCGC-4983/2020|EPI_ISL_621814|2020 .................... .................... .................... .................... .................... ....................
P 9355 hCoV-19/Denmark/DCGC-5001/2020|EPI_ISL_621816|2020 .................... .................... .................... .................... .................... ....................
P 9356 hCoV-19/Denmark/DCGC-5008/2020|EPI_ISL_621817|2020 .................... .................... .................... .................... .................... ....................
P 9357 hCoV-19/Denmark/DCGC-5014/2020|EPI_ISL_621818|2020 .................... .................... .................... .................... .................... ....................
P 9358 hCoV-19/Denmark/DCGC-5027/2020|EPI_ISL_621819|2020 .................... .................... .................... .................... .................... ....................
P 9359 hCoV-19/Denmark/DCGC-5033/2020|EPI_ISL_621820|2020 .................... .................... .................... .................... .................... ....................
P 9360 hCoV-19/Denmark/DCGC-5039/2020|EPI_ISL_621821|2020 .................... .................... .................... .................... .................... ....................
P 9361 hCoV-19/Denmark/DCGC-5045/2020|EPI_ISL_621822|2020 .................... .................... .................... .................... .................... ....................
P 9362 hCoV-19/Denmark/DCGC-6338/2020|EPI_ISL_621823|2020 .................... .A.................. .................... .................... .................... ....................
P 9363 hCoV-19/Denmark/DCGC-1721/2020|EPI_ISL_621824|2020 .................... .................... .................... .................... .................... ....................
P 9364 hCoV-19/Denmark/DCGC-1671/2020|EPI_ISL_621825|2020 .................... .................... .................... .................... .................... ....................
P 9365 hCoV-19/Denmark/DCGC-5225/2020|EPI_ISL_621826|2020 .................... .................... .................... .................... .................... ....................
P 9366 hCoV-19/Denmark/DCGC-4406/2020|EPI_ISL_621827|2020 .................... .................... .................... .................... .................... ....................
P 9367 hCoV-19/Denmark/DCGC-4407/2020|EPI_ISL_621828|2020 .................... .................... .................... .................... .................... ....................
P 9368 hCoV-19/Denmark/DCGC-4408/2020|EPI_ISL_621829|2020 .................... .................... .................... .................... .................... ....................
P 9369 hCoV-19/Denmark/DCGC-4409/2020|EPI_ISL_621830|2020 .................... .................... .................... .................... .................... ....................
P 9370 hCoV-19/Denmark/DCGC-4411/2020|EPI_ISL_621832|2020 .................... .................... .................... .................... .................... ....................
P 9371 hCoV-19/Denmark/DCGC-4412/2020|EPI_ISL_621833|2020 .................... .................... .................... .................... .................... ....................
P 9372 hCoV-19/Denmark/DCGC-4413/2020|EPI_ISL_621834|2020 .................... .................... .................... .................... .................... ....................
P 9373 hCoV-19/Denmark/DCGC-4414/2020|EPI_ISL_621835|2020 .................... .................... .................... .................... .................... ....................
P 9374 hCoV-19/Denmark/DCGC-4415/2020|EPI_ISL_621836|2020 .................... .................... .................... .................... .................... ....................
P 9375 hCoV-19/Denmark/DCGC-4416/2020|EPI_ISL_621837|2020 .................... .................... .................... .................... .................... ....................
P 9376 hCoV-19/Denmark/DCGC-4417/2020|EPI_ISL_621838|2020 .................... .................... .................... .................... .................... ....................
P 9377 hCoV-19/Denmark/DCGC-4418/2020|EPI_ISL_621839|2020 .................... .................... .................... .................... .................... ....................
P 9378 hCoV-19/Denmark/DCGC-4419/2020|EPI_ISL_621840|2020 .................... .................... .................... .................... .................... ....................
P 9379 hCoV-19/Denmark/DCGC-4420/2020|EPI_ISL_621841|2020 .................... .................... .................... .................... .................... ....................
P 9380 hCoV-19/Denmark/DCGC-4421/2020|EPI_ISL_621842|2020 .................... .................... .................... .................... .................... ....................
P 9381 hCoV-19/England/ALDP-AA125C/2020|EPI_ISL_624040|20 .................... .................... .................... .................... .................... ....................
P 9382 hCoV-19/England/ALDP-AA0EC2/2020|EPI_ISL_624041|20 .................... .................... .................... .................... .................... ....................
P 9383 hCoV-19/England/QEUH-AAEED3/2020|EPI_ISL_624043|20 .................... .................... .................... .................... .................... ....................
P 9384 hCoV-19/England/ALDP-AA1993/2020|EPI_ISL_624047|20 .................... .................... .................... .................... .................... ....................
P 9385 hCoV-19/England/ALDP-AA412F/2020|EPI_ISL_624048|20 .................... .................... .................... .................... .................... ....................
P 9386 hCoV-19/Scotland/QEUH-AAE51A/2020|EPI_ISL_624049|2 .................... .................... .................... .................... .................... ....................
P 9387 hCoV-19/England/ALDP-AA2D87/2020|EPI_ISL_624050|20 .................... .A.................. .................... .................... .................... ....................
P 9388 hCoV-19/England/ALDP-AA173F/2020|EPI_ISL_624051|20 .................... .................... .................... .................... .................... ....................
P 9389 hCoV-19/England/ALDP-AA32F0/2020|EPI_ISL_624052|20 .................... .................... .................... .................... .................... ....................
P 9390 hCoV-19/England/ALDP-AA5258/2020|EPI_ISL_624053|20 .................... .................... .................... .................... .................... ....................
P 9391 hCoV-19/England/ALDP-AA2F09/2020|EPI_ISL_624054|20 .................... .................... .................... .................... .................... ....................
P 9392 hCoV-19/England/ALDP-AA14ED/2020|EPI_ISL_624055|20 .................... .................... .................... .................... .................... ....................
P 9393 hCoV-19/England/ALDP-AA41C5/2020|EPI_ISL_624056|20 .................... .................... .................... .................... .................... ....................
P 9394 hCoV-19/England/ALDP-AA2D5A/2020|EPI_ISL_624058|20 .................... .................... .................... .................... .................... ....................
P 9395 hCoV-19/England/ALDP-AA4189/2020|EPI_ISL_624059|20 .................... .................... .................... .................... .................... ....................
P 9396 hCoV-19/England/ALDP-AA1492/2020|EPI_ISL_624060|20 .................... .................... .................... .................... .................... ....................
```

| | | | | | | | | |
|---|---|---|---|---|---|---|---|---|
| P | 9397 | hCoV-19/England/ALDP-AA41F2/2020\|EPI_ISL_624062\|20 | ···················· | ···················· | ····················· | ···················· | ···················· | ···················· |
| P | 9398 | hCoV-19/England/ALDP-AA1289/2020\|EPI_ISL_624063\|20 | ···················· | ···················· | ····················· | ···················· | ···················· | ···················· |
| P | 9399 | hCoV-19/England/ALDP-AA1465/2020\|EPI_ISL_624064\|20 | ···················· | ···················· | ····················· | ···················· | ···················· | ···················· |
| P | 9400 | hCoV-19/England/ALDP-AA371F/2020\|EPI_ISL_624066\|20 | ···················· | ···················· | ····················· | ···················· | ···················· | ···················· |
| P | 9401 | hCoV-19/Scotland/QEUH-AAE592/2020\|EPI_ISL_624067\|2 | ···················· | ···················· | ····················· | ···················· | ···················· | ···················· |
| P | 9402 | hCoV-19/England/ALDP-AA0F1A/2020\|EPI_ISL_624068\|20 | ···················· | ·A·················· | ····················· | ···················· | ···················· | ···················· |
| P | 9403 | hCoV-19/England/ALDP-AA0D7A/2020\|EPI_ISL_624069\|20 | ···················· | ···················· | ····················· | ···················· | ···················· | ···················· |
| P | 9404 | hCoV-19/England/ALDP-AA31F3/2020\|EPI_ISL_624070\|20 | ···················· | ···················· | ····················· | ···················· | ···················· | ···················· |
| P | 9405 | hCoV-19/England/ALDP-AA3375/2020\|EPI_ISL_624072\|20 | ···················· | ···················· | ····················· | ···················· | ···················· | ···················· |
| P | 9406 | hCoV-19/England/QEUH-AAE41D/2020\|EPI_ISL_624073\|20 | ···················· | ···················· | ····················· | ···················· | ···················· | ···················· |
| P | 9407 | hCoV-19/England/ALDP-AA36E5/2020\|EPI_ISL_624074\|20 | ···················· | ···················· | ····················· | ···················· | ···················· | ···················· |
| P | 9408 | hCoV-19/England/ALDP-AA1377/2020\|EPI_ISL_624075\|20 | ···················· | ···················· | ····················· | ···················· | ···················· | ···················· |
| P | 9409 | hCoV-19/England/ALDP-AA3515/2020\|EPI_ISL_624076\|20 | ···················· | ···················· | ····················· | ···················· | ···················· | ···················· |
| P | 9410 | hCoV-19/England/QEUH-AAD36C/2020\|EPI_ISL_624077\|20 | ···················· | ···················· | ····················· | ···················· | ···················· | ···················· |
| P | 9411 | hCoV-19/England/ALDP-AA3357/2020\|EPI_ISL_624080\|20 | ···················· | ·A·················· | ····················· | ···················· | ···················· | ··A················· |
| P | 9412 | hCoV-19/England/ALDP-AA1298/2020\|EPI_ISL_624082\|20 | ···················· | ···················· | ····················· | ···················· | ···················· | ···················· |
| P | 9413 | hCoV-19/England/ALDP-AA376A/2020\|EPI_ISL_624083\|20 | ···················· | ···················· | ····················· | ···················· | ···················· | ···················· |
| P | 9414 | hCoV-19/England/ALDP-AA332A/2020\|EPI_ISL_624084\|20 | ···················· | ···················· | ····················· | ···················· | ···················· | ···················· |
| P | 9415 | hCoV-19/England/ALDP-AA0D6B/2020\|EPI_ISL_624085\|20 | ···················· | ···················· | ····················· | ···················· | ···················· | ···················· |
| P | 9416 | hCoV-19/England/ALDP-AA0D89/2020\|EPI_ISL_624086\|20 | ···················· | ···················· | ····················· | ···················· | ···················· | ···················· |
| P | 9417 | hCoV-19/England/ALDP-AA3551/2020\|EPI_ISL_624087\|20 | ···················· | ···················· | ····················· | ···················· | ···················· | ···················· |
| P | 9418 | hCoV-19/England/ALDP-AA2B50/2020\|EPI_ISL_624088\|20 | ···················· | ···················· | ····················· | ···················· | ···················· | ···················· |
| P | 9419 | hCoV-19/England/ALDP-AA1201/2020\|EPI_ISL_624089\|20 | ···················· | ···················· | ····················· | ···················· | ···················· | ···················· |
| P | 9420 | hCoV-19/England/ALDP-AA0DD4/2020\|EPI_ISL_624090\|20 | ···················· | ···················· | ····················· | ···················· | ···················· | ···················· |
| P | 9421 | hCoV-19/England/ALDP-AA2B14/2020\|EPI_ISL_624091\|20 | ···················· | ···················· | ····················· | ···················· | ···················· | ···················· |
| P | 9422 | hCoV-19/England/QEUH-AADC16/2020\|EPI_ISL_624092\|20 | ···················· | ···················· | ····················· | ···················· | ······T············· | ···················· |
| P | 9423 | hCoV-19/England/ALDP-AA520D/2020\|EPI_ISL_624093\|20 | ···················· | ···················· | ····················· | ···················· | ···················· | ···················· |
| P | 9424 | hCoV-19/England/ALDP-AA1386/2020\|EPI_ISL_624096\|20 | ···················· | ···················· | ····················· | ···················· | ···················· | ···················· |
| P | 9425 | hCoV-19/England/ALDP-AA56B6/2020\|EPI_ISL_624097\|20 | ···················· | ·A·················· | ····················· | ···················· | ···················· | ··A················· |
| P | 9426 | hCoV-19/England/ALDP-AA076D/2020\|EPI_ISL_624098\|20 | ···················· | ···················· | ····················· | ···················· | ···················· | ···················· |
| P | 9427 | hCoV-19/England/ALDP-AA140B/2020\|EPI_ISL_624099\|20 | ···················· | ···················· | ····················· | ···················· | ···················· | ···················· |
| P | 9428 | hCoV-19/Wales/CAMC-A81DBB/2020\|EPI_ISL_625308\|2020 | ···················· | ···················· | ····················· | ···················· | ···················· | ···················· |
| P | 9429 | hCoV-19/Scotland/CAMC-A81C81/2020\|EPI_ISL_625309\|2 | ···················· | ···················· | ····················· | ···················· | ···················· | ···················· |
| P | 9430 | hCoV-19/Wales/CAMC-A80E04/2020\|EPI_ISL_625310\|2020 | ···················· | ···················· | ····················· | ···················· | ···················· | ···················· |
| P | 9431 | hCoV-19/Wales/CAMC-A82EC6/2020\|EPI_ISL_625311\|2020 | ···················· | ···················· | ····················· | ···················· | ···················· | ···················· |
| P | 9432 | hCoV-19/Scotland/CAMC-A82084/2020\|EPI_ISL_625313\|2 | ···················· | ···················· | ····················· | ···················· | ···················· | ···················· |
| P | 9433 | hCoV-19/Wales/CAMC-A82479/2020\|EPI_ISL_625314\|2020 | ···················· | ···················· | ····················· | ···················· | ···················· | ···················· |
| P | 9434 | hCoV-19/Scotland/QEUH-A9EE03/2020\|EPI_ISL_625317\|2 | ···················· | ···················· | ····················· | ···················· | ···················· | ···················· |
| P | 9435 | hCoV-19/Scotland/QEUH-A9E98A/2020\|EPI_ISL_625319\|2 | ···················· | ···················· | ····················· | ···················· | ···················· | ···················· |
| P | 9436 | hCoV-19/Scotland/QEUH-A9EDCA/2020\|EPI_ISL_625320\|2 | ···················· | ···················· | ····················· | ···················· | ···················· | ···················· |
| P | 9437 | hCoV-19/Scotland/QEUH-A9EA3C/2020\|EPI_ISL_625321\|2 | ···················· | ···················· | ····················· | ···················· | ···················· | ···················· |
| P | 9438 | hCoV-19/Scotland/QEUH-A9B352/2020\|EPI_ISL_625322\|2 | ···················· | ···················· | ····················· | ···················· | ···················· | ···················· |
| P | 9439 | hCoV-19/Scotland/QEUH-A9EA0F/2020\|EPI_ISL_625323\|2 | ···················· | ···················· | ····················· | ···················· | ···················· | ···················· |
| P | 9440 | hCoV-19/Wales/QEUH-A9FFE1/2020\|EPI_ISL_625324\|2020 | ···················· | ···················· | ····················· | ···················· | ···················· | ···················· |
| P | 9441 | hCoV-19/Scotland/QEUH-A9EBC0/2020\|EPI_ISL_625325\|2 | ···················· | ···················· | ····················· | ···················· | ···················· | ···················· |
| P | 9442 | hCoV-19/Scotland/QEUH-A9EBA2/2020\|EPI_ISL_625327\|2 | ···················· | ···················· | ····················· | ···················· | ···················· | ···················· |
| P | 9443 | hCoV-19/Scotland/QEUH-A9B5D4/2020\|EPI_ISL_625329\|2 | ···················· | ···················· | ····················· | ···················· | ···················· | ···················· |
| P | 9444 | hCoV-19/Scotland/QEUH-A9EA96/2020\|EPI_ISL_625370\|2 | ···················· | ···················· | ····················· | ···················· | ···················· | ···················· |
| P | 9445 | hCoV-19/Scotland/QEUH-A9B5F2/2020\|EPI_ISL_625371\|2 | ···················· | ·A·················· | ····················· | ···················· | ···················· | ···················· |
| P | 9446 | hCoV-19/Scotland/QEUH-A9EA78/2020\|EPI_ISL_625372\|2 | ···················· | ···················· | ····················· | ···················· | ···················· | ···················· |
| P | 9447 | hCoV-19/Scotland/QEUH-A9B3AD/2020\|EPI_ISL_625373\|2 | ···················· | ···················· | ····················· | ···················· | ···················· | ···················· |
| P | 9448 | hCoV-19/Scotland/QEUH-A9E9A8/2020\|EPI_ISL_625374\|2 | ···················· | ···················· | ····················· | ···················· | ···················· | ···················· |
| P | 9449 | hCoV-19/Scotland/QEUH-A9F057/2020\|EPI_ISL_625375\|2 | ···················· | ···················· | ····················· | ···················· | ···················· | ···················· |
| P | 9450 | hCoV-19/Scotland/QEUH-A9EE4F/2020\|EPI_ISL_625377\|2 | ···················· | ···················· | ····················· | ···················· | ···················· | ···················· |
| P | 9451 | hCoV-19/Scotland/QEUH-A9B932/2020\|EPI_ISL_625378\|2 | ···················· | ···················· | ····················· | ···················· | ···················· | ···················· |
| P | 9452 | hCoV-19/Scotland/QEUH-A9E805/2020\|EPI_ISL_625379\|2 | ···················· | ···················· | ····················· | ···················· | ···················· | ···················· |
| P | 9453 | hCoV-19/Scotland/QEUH-A9F0DF/2020\|EPI_ISL_625380\|2 | ···················· | ···················· | ····················· | ···················· | ···················· | ···················· |
| P | 9454 | hCoV-19/Scotland/QEUH-A9EA87/2020\|EPI_ISL_625381\|2 | ···················· | ···················· | ····················· | ···················· | ···················· | ···················· |
| P | 9455 | hCoV-19/Wales/QEUH-AA0509/2020\|EPI_ISL_625382\|2020 | ···················· | ···················· | ····················· | ···················· | ···················· | ···················· |
| P | 9456 | hCoV-19/Scotland/QEUH-A9F0FD/2020\|EPI_ISL_625383\|2 | ···················· | ···················· | ····················· | ···················· | ···················· | ···················· |
| P | 9457 | hCoV-19/Scotland/QEUH-A9AE61/2020\|EPI_ISL_625384\|2 | ···················· | ···················· | ····················· | ···················· | ···················· | ···················· |
| P | 9458 | hCoV-19/Scotland/QEUH-A9AE07/2020\|EPI_ISL_625385\|2 | ···················· | ···················· | ····················· | ···················· | ···················· | ···················· |
| P | 9459 | hCoV-19/Scotland/QEUH-A9AE8F/2020\|EPI_ISL_625386\|2 | ···················· | ···················· | ····················· | ···················· | ···················· | ···················· |
| P | 9460 | hCoV-19/Scotland/QEUH-A9B3DA/2020\|EPI_ISL_625387\|2 | ···················· | ···················· | ····················· | ···················· | ···················· | ···················· |
| P | 9461 | hCoV-19/Scotland/QEUH-A9B5E3/2020\|EPI_ISL_625388\|2 | ···················· | ···················· | ····················· | ···················· | ···················· | ···················· |
| P | 9462 | hCoV-19/Wales/QEUH-A9F4A6/2020\|EPI_ISL_625389\|2020 | ···················· | ···················· | ····················· | ···················· | ···················· | ···················· |
| P | 9463 | hCoV-19/Scotland/QEUH-A9B431/2020\|EPI_ISL_625390\|2 | ···················· | ···················· | ····················· | ···················· | ···················· | ···················· |
| P | 9464 | hCoV-19/Scotland/QEUH-A9E9C6/2020\|EPI_ISL_625391\|2 | ···················· | ···················· | ····················· | ···················· | ···················· | ···················· |
| P | 9465 | hCoV-19/Scotland/QEUH-A9B361/2020\|EPI_ISL_625392\|2 | ···················· | ···················· | ····················· | ···················· | ···················· | ···················· |
| P | 9466 | hCoV-19/Scotland/QEUH-A9B60E/2020\|EPI_ISL_625393\|2 | ···················· | ···················· | ····················· | ···················· | ···················· | ···················· |
| P | 9467 | hCoV-19/Scotland/QEUH-A9B48C/2020\|EPI_ISL_625394\|2 | ···················· | ···················· | ····················· | ···················· | ···················· | ···················· |
| P | 9468 | hCoV-19/Scotland/QEUH-A9B659/2020\|EPI_ISL_625395\|2 | ···················· | ···················· | ····················· | ···················· | ···················· | ···················· |
| P | 9469 | hCoV-19/Wales/ALDP-A87D3D/2020\|EPI_ISL_625396\|2020 | ···················· | ···················· | ····················· | ···················· | ···················· | ···················· |
| P | 9470 | hCoV-19/Wales/ALDP-A86369/2020\|EPI_ISL_625397\|2020 | ···················· | ···················· | ····················· | ···················· | ···················· | ···················· |
| P | 9471 | hCoV-19/Wales/ALDP-A8640C/2020\|EPI_ISL_625398\|2020 | ···················· | ···················· | ····················· | ···················· | ···················· | ···················· |
| P | 9472 | hCoV-19/Wales/ALDP-A862D5/2020\|EPI_ISL_625399\|2020 | ···················· | ···················· | ····················· | ···················· | ···················· | ···················· |
| P | 9473 | hCoV-19/Denmark/DCGC-1102/2020\|EPI_ISL_614600\|2020 | ···················· | ···················· | ····················· | ···················· | ···················· | ···················· |
| P | 9474 | hCoV-19/Denmark/DCGC-1121/2020\|EPI_ISL_614601\|2020 | ···················· | ···················· | ····················· | ···················· | ···················· | ···················· |
| P | 9475 | hCoV-19/Denmark/DCGC-1095/2020\|EPI_ISL_614602\|2020 | ···················· | ···················· | ····················· | ···················· | ···················· | ···················· |
| P | 9476 | hCoV-19/Denmark/DCGC-1112/2020\|EPI_ISL_614603\|2020 | ···················· | ···················· | ····················· | ···················· | ···················· | ···················· |

| | | | | | | | | |
|---|---|---|---|---|---|---|---|---|
| P | 9477 | hCoV-19/Denmark/DCGC-1117/2020\|EPI_ISL_614604\|2020 | .................... | .................... | .................... | .................... | .................... | .................... |
| P | 9478 | hCoV-19/Denmark/DCGC-1124/2020\|EPI_ISL_614605\|2020 | .................... | .................... | .................... | .................... | .................... | .................... |
| P | 9479 | hCoV-19/Denmark/DCGC-1137/2020\|EPI_ISL_614606\|2020 | .................... | .................... | .................... | .................... | .................... | .................... |
| P | 9480 | hCoV-19/Denmark/DCGC-1092/2020\|EPI_ISL_614607\|2020 | .................... | .................... | .................... | .................... | .................... | .................... |
| P | 9481 | hCoV-19/Denmark/DCGC-1096/2020\|EPI_ISL_614608\|2020 | .................... | .................... | .................... | .................... | .................... | .................... |
| P | 9482 | hCoV-19/Denmark/DCGC-1125/2020\|EPI_ISL_614609\|2020 | .................... | .................... | .................... | .................... | .................... | .................... |
| P | 9483 | hCoV-19/Denmark/DCGC-1129/2020\|EPI_ISL_614610\|2020 | .................... | .................... | .................... | .................... | .................... | .................... |
| P | 9484 | hCoV-19/Denmark/DCGC-1133/2020\|EPI_ISL_614611\|2020 | .................... | .................... | .................... | .................... | .................... | .................... |
| P | 9485 | hCoV-19/Denmark/DCGC-1103/2020\|EPI_ISL_614612\|2020 | .................... | .................... | .................... | .................... | .................... | .................... |
| P | 9486 | hCoV-19/Denmark/DCGC-1118/2020\|EPI_ISL_614613\|2020 | .................... | .................... | .................... | .................... | .................... | .................... |
| P | 9487 | hCoV-19/Denmark/DCGC-1134/2020\|EPI_ISL_614614\|2020 | .................... | .................... | .................... | .................... | .................... | .................... |
| P | 9488 | hCoV-19/Denmark/DCGC-1093/2020\|EPI_ISL_614615\|2020 | .................... | .................... | .................... | .................... | .................... | .................... |
| P | 9489 | hCoV-19/Denmark/DCGC-1098/2020\|EPI_ISL_614616\|2020 | .................... | .................... | .................... | .................... | .................... | .................... |
| P | 9490 | hCoV-19/Denmark/DCGC-1107/2020\|EPI_ISL_614617\|2020 | .................... | .................... | .................... | .................... | .................... | .................... |
| P | 9491 | hCoV-19/Denmark/DCGC-1130/2020\|EPI_ISL_614618\|2020 | .................... | .................... | .................... | .................... | .................... | .................... |
| P | 9492 | hCoV-19/Denmark/DCGC-1135/2020\|EPI_ISL_614619\|2020 | .................... | .................... | .................... | .................... | .................... | .................... |
| P | 9493 | hCoV-19/Denmark/DCGC-1099/2020\|EPI_ISL_614620\|2020 | .................... | .................... | .................... | .................... | .................... | .................... |
| P | 9494 | hCoV-19/Denmark/DCGC-1104/2020\|EPI_ISL_614621\|2020 | .................... | .................... | .................... | .................... | .................... | .................... |
| P | 9495 | hCoV-19/Denmark/DCGC-1113/2020\|EPI_ISL_614622\|2020 | .................... | .................... | .................... | .................... | .................... | .................... |
| P | 9496 | hCoV-19/Denmark/DCGC-1119/2020\|EPI_ISL_614623\|2020 | .................... | .................... | .................... | .................... | .................... | .................... |
| P | 9497 | hCoV-19/Denmark/DCGC-1122/2020\|EPI_ISL_614624\|2020 | .................... | .................... | .................... | .................... | .................... | .................... |
| P | 9498 | hCoV-19/Denmark/DCGC-1136/2020\|EPI_ISL_614625\|2020 | .................... | .................... | .................... | .................... | .................... | .................... |
| P | 9499 | hCoV-19/Denmark/DCGC-1146/2020\|EPI_ISL_614626\|2020 | .................... | .................... | .................... | .................... | .................... | .................... |
| P | 9500 | hCoV-19/Denmark/DCGC-1155/2020\|EPI_ISL_614627\|2020 | .................... | .................... | .................... | .................... | .................... | .................... |
| P | 9501 | hCoV-19/Denmark/DCGC-1160/2020\|EPI_ISL_614628\|2020 | .................... | .................... | .................... | .................... | .................... | .................... |
| P | 9502 | hCoV-19/Denmark/DCGC-1165/2020\|EPI_ISL_614629\|2020 | .................... | .................... | .................... | .................... | .................... | .................... |
| P | 9503 | hCoV-19/Denmark/DCGC-1171/2020\|EPI_ISL_614630\|2020 | .................... | .................... | .................... | .................... | .................... | .................... |
| P | 9504 | hCoV-19/Denmark/DCGC-919/2020\|EPI_ISL_614631\|2020- | .................... | .................... | .................... | .................... | .................... | .................... |
| P | 9505 | hCoV-19/Denmark/DCGC-1138/2020\|EPI_ISL_614632\|2020 | .................... | .................... | .................... | .................... | .................... | .................... |
| P | 9506 | hCoV-19/Denmark/DCGC-1141/2020\|EPI_ISL_614633\|2020 | .................... | .................... | .................... | .................... | .................... | .................... |
| P | 9507 | hCoV-19/Denmark/DCGC-1145/2020\|EPI_ISL_614634\|2020 | .................... | .................... | .................... | .................... | .................... | .................... |
| P | 9508 | hCoV-19/Denmark/DCGC-1147/2020\|EPI_ISL_614635\|2020 | .................... | .................... | .................... | .................... | .................... | .................... |
| P | 9509 | hCoV-19/Denmark/DCGC-1150/2020\|EPI_ISL_614636\|2020 | .................... | .................... | .................... | .................... | .................... | .................... |
| P | 9510 | hCoV-19/Denmark/DCGC-1161/2020\|EPI_ISL_614637\|2020 | .................... | .................... | .................... | .................... | .................... | .................... |
| P | 9511 | hCoV-19/Denmark/DCGC-5229/2020\|EPI_ISL_615900\|2020 | ........T........... | .................... | .................... | .................... | .................... | .................... |
| P | 9512 | hCoV-19/Denmark/DCGC-5230/2020\|EPI_ISL_615901\|2020 | .................... | .................... | .................... | .................... | .................... | .................... |
| P | 9513 | hCoV-19/Denmark/DCGC-5231/2020\|EPI_ISL_615902\|2020 | .................... | .................... | .................... | .................... | .................... | .................... |
| P | 9514 | hCoV-19/Denmark/DCGC-5232/2020\|EPI_ISL_615903\|2020 | .................... | .................... | .................... | .................... | .................... | .................... |
| P | 9515 | hCoV-19/Denmark/DCGC-5233/2020\|EPI_ISL_615904\|2020 | .................... | .................... | .................... | .................... | .................... | .................... |
| P | 9516 | hCoV-19/Denmark/DCGC-5234/2020\|EPI_ISL_615905\|2020 | .................... | .................... | .................... | .................... | .................... | .................... |
| P | 9517 | hCoV-19/Denmark/DCGC-3579/2020\|EPI_ISL_615907\|2020 | .................... | .................... | .................... | .................... | .................... | .................... |
| P | 9518 | hCoV-19/Denmark/DCGC-3580/2020\|EPI_ISL_615908\|2020 | .................... | .................... | .................... | .................... | .................... | .................... |
| P | 9519 | hCoV-19/Denmark/DCGC-3581/2020\|EPI_ISL_615909\|2020 | .................... | .................... | .................... | .................... | .................... | .................... |
| P | 9520 | hCoV-19/Denmark/DCGC-3582/2020\|EPI_ISL_615910\|2020 | .................... | .................... | .................... | .................... | .................... | .................... |
| P | 9521 | hCoV-19/Denmark/DCGC-3583/2020\|EPI_ISL_615911\|2020 | .................... | .................... | .................... | .................... | .................... | .................... |
| P | 9522 | hCoV-19/Denmark/DCGC-3096/2020\|EPI_ISL_615912\|2020 | .................... | .................... | .................... | .................... | .................... | .................... |
| P | 9523 | hCoV-19/Denmark/DCGC-3097/2020\|EPI_ISL_615913\|2020 | .................... | .................... | .................... | .................... | .................... | .................... |
| P | 9524 | hCoV-19/Denmark/DCGC-3104/2020\|EPI_ISL_615920\|2020 | .................... | .................... | .................... | .................... | .................... | .................... |
| P | 9525 | hCoV-19/Denmark/DCGC-3105/2020\|EPI_ISL_615921\|2020 | .................... | .................... | .................... | .................... | .................... | .................... |
| P | 9526 | hCoV-19/Denmark/DCGC-3247/2020\|EPI_ISL_615923\|2020 | .................... | .................... | .................... | .................... | .................... | .................... |
| P | 9527 | hCoV-19/Denmark/DCGC-3108/2020\|EPI_ISL_615925\|2020 | .................... | .................... | .................... | .................... | .................... | .................... |
| P | 9528 | hCoV-19/Denmark/DCGC-3109/2020\|EPI_ISL_615926\|2020 | .................... | .................... | .................... | .................... | .................... | .................... |
| P | 9529 | hCoV-19/Denmark/DCGC-3110/2020\|EPI_ISL_615927\|2020 | .................... | .................... | .................... | .................... | .................... | .................... |
| P | 9530 | hCoV-19/Denmark/DCGC-3111/2020\|EPI_ISL_615928\|2020 | .................... | .................... | .................... | .................... | .................... | .................... |
| P | 9531 | hCoV-19/Denmark/DCGC-3112/2020\|EPI_ISL_615929\|2020 | .................... | .................... | .................... | .................... | .................... | .................... |
| P | 9532 | hCoV-19/Denmark/DCGC-3113/2020\|EPI_ISL_615930\|2020 | .................... | .................... | .................... | .................... | .................... | .................... |
| P | 9533 | hCoV-19/Denmark/DCGC-3114/2020\|EPI_ISL_615931\|2020 | .................... | .................... | .................... | .................... | .................... | .................... |
| P | 9534 | hCoV-19/Denmark/DCGC-3115/2020\|EPI_ISL_615932\|2020 | .................... | .................... | .................... | .................... | .................... | .................... |
| P | 9535 | hCoV-19/Denmark/DCGC-3116/2020\|EPI_ISL_615933\|2020 | .................... | .................... | .................... | .................... | .................... | .................... |
| P | 9536 | hCoV-19/Denmark/DCGC-3117/2020\|EPI_ISL_615934\|2020 | .................... | .................... | .................... | .................... | .................... | .................... |
| P | 9537 | hCoV-19/Denmark/DCGC-3120/2020\|EPI_ISL_615937\|2020 | .................... | .................... | .................... | .................... | .................... | .................... |
| P | 9538 | hCoV-19/Denmark/DCGC-3121/2020\|EPI_ISL_615938\|2020 | .................... | .................... | .................... | .................... | .................... | .................... |
| P | 9539 | hCoV-19/Denmark/DCGC-3122/2020\|EPI_ISL_615939\|2020 | .................... | .................... | .................... | .................... | .................... | .................... |
| P | 9540 | hCoV-19/Denmark/DCGC-3123/2020\|EPI_ISL_615940\|2020 | .................... | .................... | .................... | .................... | .................... | .................... |
| P | 9541 | hCoV-19/Denmark/DCGC-3124/2020\|EPI_ISL_615941\|2020 | .................... | .................... | .................... | .................... | .................... | .................... |
| P | 9542 | hCoV-19/Denmark/DCGC-3125/2020\|EPI_ISL_615942\|2020 | .................... | .................... | .................... | .................... | .................... | .................... |
| P | 9543 | hCoV-19/Denmark/DCGC-3248/2020\|EPI_ISL_615943\|2020 | .................... | .................... | .................... | .................... | .................... | .................... |
| P | 9544 | hCoV-19/Denmark/DCGC-3126/2020\|EPI_ISL_615944\|2020 | .................... | .................... | .................... | .................... | .................... | .................... |
| P | 9545 | hCoV-19/Denmark/DCGC-3127/2020\|EPI_ISL_615945\|2020 | .................... | .................... | .................... | .................... | .................... | .................... |
| P | 9546 | hCoV-19/Denmark/DCGC-3249/2020\|EPI_ISL_615946\|2020 | .................... | .................... | .................... | .................... | .................... | .................... |
| P | 9547 | hCoV-19/Denmark/DCGC-3128/2020\|EPI_ISL_615947\|2020 | .................... | .................... | .................... | .................... | .................... | .................... |
| P | 9548 | hCoV-19/Denmark/DCGC-3129/2020\|EPI_ISL_615948\|2020 | .................... | .................... | .................... | .................... | .................... | .................... |
| P | 9549 | hCoV-19/Denmark/DCGC-3130/2020\|EPI_ISL_615949\|2020 | .................... | .................... | .................... | .................... | .................... | .................... |
| P | 9550 | hCoV-19/Denmark/DCGC-3131/2020\|EPI_ISL_615950\|2020 | .................... | .................... | .................... | .................... | .................... | .................... |
| P | 9551 | hCoV-19/Denmark/DCGC-3132/2020\|EPI_ISL_615951\|2020 | .................... | .................... | .................... | .................... | .................... | .................... |
| P | 9552 | hCoV-19/Denmark/DCGC-3133/2020\|EPI_ISL_615952\|2020 | .................... | .................... | .................... | .................... | .................... | .................... |
| P | 9553 | hCoV-19/Denmark/DCGC-3134/2020\|EPI_ISL_615953\|2020 | .................... | .................... | .................... | .................... | .................... | .................... |
| P | 9554 | hCoV-19/Denmark/DCGC-3135/2020\|EPI_ISL_615954\|2020 | .................... | .................... | .................... | .................... | .................... | .................... |
| P | 9555 | hCoV-19/Denmark/DCGC-3136/2020\|EPI_ISL_615955\|2020 | .................... | .................... | .................... | .................... | .................... | .................... |
| P | 9556 | hCoV-19/Denmark/DCGC-3137/2020\|EPI_ISL_615956\|2020 | .................... | .................... | .................... | .................... | .................... | .................... |

P 9557 hCoV-19/Denmark/DCGC-3138/2020|EPI_ISL_615957|2020 ···················· ···················· ····················· ···················· ···················· ····················
P 9558 hCoV-19/Denmark/DCGC-3140/2020|EPI_ISL_615959|2020 ···················· ···················· ····················· ···················· ···················· ····················
P 9559 hCoV-19/Denmark/DCGC-3141/2020|EPI_ISL_615960|2020 ···················· ···················· ····················· ···················· ···················· ····················
P 9560 hCoV-19/Denmark/DCGC-3250/2020|EPI_ISL_615961|2020 ···················· ···················· ····················· ···················· ···················· ····················
P 9561 hCoV-19/Denmark/DCGC-3142/2020|EPI_ISL_615962|2020 ···················· ···················· ····················· ···················· ···················· ····················
P 9562 hCoV-19/Denmark/DCGC-3145/2020|EPI_ISL_615965|2020 ···················· ···················· ····················· ···················· ···················· ····················
P 9563 hCoV-19/Denmark/DCGC-3146/2020|EPI_ISL_615966|2020 ···················· ···················· ····················· ···················· ···················· ····················
P 9564 hCoV-19/Denmark/DCGC-3147/2020|EPI_ISL_615967|2020 ···················· ···················· ····················· ···················· ···················· ····················
P 9565 hCoV-19/Denmark/DCGC-3148/2020|EPI_ISL_615968|2020 ···················· ···················· ····················· ···················· ···················· ····················
P 9566 hCoV-19/Denmark/DCGC-1482/2020|EPI_ISL_618100|2020 ···················· ···················· ····················· ···················· ···················· ····················
P 9567 hCoV-19/Denmark/DCGC-1481/2020|EPI_ISL_618101|2020 ···················· ···················· ····················· ···················· ···················· ····················
P 9568 hCoV-19/Denmark/DCGC-1480/2020|EPI_ISL_618102|2020 ···················· ···················· ····················· ···················· ···················· ····················
P 9569 hCoV-19/Denmark/DCGC-1479/2020|EPI_ISL_618103|2020 ···················· ···················· ····················· ···················· ···················· ····················
P 9570 hCoV-19/Denmark/DCGC-1478/2020|EPI_ISL_618104|2020 ···················· ···················· ····················· ···················· ···················· ····················
P 9571 hCoV-19/Denmark/DCGC-1502/2020|EPI_ISL_618105|2020 ···················· ···················· ····················· ···················· ···················· ····················
P 9572 hCoV-19/Denmark/DCGC-1505/2020|EPI_ISL_618108|2020 ···················· ···················· ····················· ···················· ···················· ····················
P 9573 hCoV-19/Denmark/DCGC-1506/2020|EPI_ISL_618109|2020 ···················· ···················· ····················· ···················· ···················· ····················
P 9574 hCoV-19/Denmark/DCGC-1507/2020|EPI_ISL_618110|2020 ···················· ···················· ····················· ···················· ···················· ····················
P 9575 hCoV-19/Denmark/DCGC-1508/2020|EPI_ISL_618111|2020 ···················· ···················· ····················· ···················· ···················· ····················
P 9576 hCoV-19/Denmark/DCGC-1509/2020|EPI_ISL_618112|2020 ···················· ···················· ····················· ···················· ···················· ····················
P 9577 hCoV-19/Denmark/DCGC-1510/2020|EPI_ISL_618113|2020 ···················· ···················· ····················· ···················· ···················· ····················
P 9578 hCoV-19/Denmark/DCGC-1511/2020|EPI_ISL_618114|2020 ···················· ···················· ····················· ···················· ···················· ····················
P 9579 hCoV-19/Denmark/DCGC-1512/2020|EPI_ISL_618115|2020 ···················· ···················· ····················· ···················· ···················· ····················
P 9580 hCoV-19/Denmark/DCGC-1513/2020|EPI_ISL_618116|2020 ···················· ···················· ····················· ···················· ···················· ····················
P 9581 hCoV-19/Denmark/DCGC-1501/2020|EPI_ISL_618117|2020 ·········T·········· ···················· ····················· ···················· ···················· ····················
P 9582 hCoV-19/Denmark/DCGC-1514/2020|EPI_ISL_618118|2020 ···················· ···················· ····················· ···················· ···················· ····················
P 9583 hCoV-19/Denmark/DCGC-1574/2020|EPI_ISL_618144|2020 ···················· ···················· ····················· ···················· ···················· ····················
P 9584 hCoV-19/Denmark/DCGC-1575/2020|EPI_ISL_618145|2020 ···················· ···················· ····················· ···················· ···················· ····················
P 9585 hCoV-19/Denmark/DCGC-1578/2020|EPI_ISL_618148|2020 ···················· ···················· ····················· ···················· ···················· ····················
P 9586 hCoV-19/Denmark/DCGC-1559/2020|EPI_ISL_618149|2020 ···················· ···················· ····················· ···················· ···················· ····················
P 9587 hCoV-19/Denmark/DCGC-1686/2020|EPI_ISL_618150|2020 ···················· ···················· ····················· ···················· ···················· ····················
P 9588 hCoV-19/Denmark/DCGC-1685/2020|EPI_ISL_618154|2020 ···················· ···················· ····················· ···················· ···················· ····················
P 9589 hCoV-19/Denmark/DCGC-2030/2020|EPI_ISL_618155|2020 ···················· ···················· ····················· ···················· ···················· ····················
P 9590 hCoV-19/Denmark/DCGC-2026/2020|EPI_ISL_618156|2020 ···················· ···················· ····················· ···················· ···················· ····················
P 9591 hCoV-19/Denmark/DCGC-1670/2020|EPI_ISL_618157|2020 ···················· ···················· ····················· ···················· ···················· ····················
P 9592 hCoV-19/Denmark/DCGC-1690/2020|EPI_ISL_618158|2020 ···················· ···················· ····················· ···················· ···················· ····················
P 9593 hCoV-19/Denmark/DCGC-1691/2020|EPI_ISL_618159|2020 ···················· ···················· ····················· ···················· ···················· ····················
P 9594 hCoV-19/Denmark/DCGC-1692/2020|EPI_ISL_618160|2020 ···················· ···················· ····················· ···················· ···················· ····················
P 9595 hCoV-19/Denmark/DCGC-1693/2020|EPI_ISL_618161|2020 ···················· ···················· ····················· ···················· ···················· ····················
P 9596 hCoV-19/Denmark/DCGC-1694/2020|EPI_ISL_618162|2020 ···················· ···················· ····················· ···················· ···················· ····················
P 9597 hCoV-19/Denmark/DCGC-1695/2020|EPI_ISL_618163|2020 ···················· ···················· ····················· ···················· ···················· ····················
P 9598 hCoV-19/Denmark/DCGC-1696/2020|EPI_ISL_618164|2020 ···················· ···················· ····················· ···················· ···················· ····················
P 9599 hCoV-19/Denmark/DCGC-1698/2020|EPI_ISL_618165|2020 ···················· ···················· ····················· ···················· ···················· ····················
P 9600 hCoV-19/Denmark/DCGC-1699/2020|EPI_ISL_618166|2020 ···················· ···················· ····················· ···················· ···················· ····················
P 9601 hCoV-19/Denmark/DCGC-1700/2020|EPI_ISL_618167|2020 ···················· ···················· ····················· ···················· ···················· ····················
P 9602 hCoV-19/Denmark/DCGC-1702/2020|EPI_ISL_618169|2020 ···················· ···················· ····················· ···················· ···················· ····················
P 9603 hCoV-19/Denmark/DCGC-1703/2020|EPI_ISL_618170|2020 ···················· ···················· ····················· ···················· ···················· ····················
P 9604 hCoV-19/Denmark/DCGC-1704/2020|EPI_ISL_618171|2020 ···················· ···················· ····················· ···················· ···················· ····················
P 9605 hCoV-19/Denmark/DCGC-1697/2020|EPI_ISL_618173|2020 ···················· ···················· ····················· ···················· ···················· ····················
P 9606 hCoV-19/Denmark/DCGC-2007/2020|EPI_ISL_618174|2020 ···················· ···················· ····················· ···················· ···················· ····················
P 9607 hCoV-19/Denmark/DCGC-1801/2020|EPI_ISL_618175|2020 ···················· ···················· ····················· ···················· ···················· ····················
P 9608 hCoV-19/Denmark/DCGC-1804/2020|EPI_ISL_618176|2020 ···················· ···················· ····················· ···················· ···················· ····················
P 9609 hCoV-19/Denmark/DCGC-1768/2020|EPI_ISL_618177|2020 ···················· ···················· ····················· ···················· ···················· ····················
P 9610 hCoV-19/Denmark/DCGC-1790/2020|EPI_ISL_618178|2020 ···················· ···················· ····················· ···················· ···················· ····················
P 9611 hCoV-19/Denmark/DCGC-1793/2020|EPI_ISL_618179|2020 ···················· ···················· ····················· ···················· ···················· ····················
P 9612 hCoV-19/Denmark/DCGC-1798/2020|EPI_ISL_618180|2020 ···················· ···················· ····················· ···················· ···················· ····················
P 9613 hCoV-19/Denmark/DCGC-1802/2020|EPI_ISL_618181|2020 ···················· ···················· ····················· ···················· ···················· ····················
P 9614 hCoV-19/Denmark/DCGC-1806/2020|EPI_ISL_618182|2020 ···················· ···················· ····················· ···················· ···················· ····················
P 9615 hCoV-19/Denmark/DCGC-1780/2020|EPI_ISL_618183|2020 ···················· ···················· ····················· ···················· ···················· ····················
P 9616 hCoV-19/Denmark/DCGC-1794/2020|EPI_ISL_618184|2020 ···················· ···················· ····················· ···················· ···················· ····················
P 9617 hCoV-19/Denmark/DCGC-1943/2020|EPI_ISL_618185|2020 ···················· ···················· ····················· ···················· ···················· ····················
P 9618 hCoV-19/Denmark/DCGC-1769/2020|EPI_ISL_618186|2020 ···················· ···················· ····················· ···················· ···················· ····················
P 9619 hCoV-19/Denmark/DCGC-1772/2020|EPI_ISL_618187|2020 ···················· ···················· ····················· ···················· ···················· ····················
P 9620 hCoV-19/Denmark/DCGC-1777/2020|EPI_ISL_618188|2020 ···················· ···················· ····················· ···················· ···················· ····················
P 9621 hCoV-19/Denmark/DCGC-1781/2020|EPI_ISL_618189|2020 ···················· ···················· ····················· ···················· ···················· ····················
P 9622 hCoV-19/Denmark/DCGC-1785/2020|EPI_ISL_618190|2020 ···················· ···················· ····················· ···················· ···················· ····················
P 9623 hCoV-19/Denmark/DCGC-1807/2020|EPI_ISL_618191|2020 ···················· ···················· ····················· ···················· ···················· ····················
P 9624 hCoV-19/Denmark/DCGC-1782/2020|EPI_ISL_618194|2020 ···················· ···················· ····················· ···················· ···················· ····················
P 9625 hCoV-19/Denmark/DCGC-1786/2020|EPI_ISL_618195|2020 ···················· ···················· ····················· ···················· ···················· ····················
P 9626 hCoV-19/Denmark/DCGC-1799/2020|EPI_ISL_618196|2020 ···················· ···················· ····················· ···················· ···················· ····················
P 9627 hCoV-19/Denmark/DCGC-1808/2020|EPI_ISL_618197|2020 ···················· ···················· ····················· ···················· ···················· ····················
P 9628 hCoV-19/Denmark/DCGC-1765/2020|EPI_ISL_618198|2020 ···················· ···················· ····················· ···················· ···················· ····················
P 9629 hCoV-19/Denmark/DCGC-1770/2020|EPI_ISL_618199|2020 ···················· ···················· ····················· ···················· ···················· ····················
P 9630 hCoV-19/Denmark/DCGC-6510/2020|EPI_ISL_619410|2020 ···················· ···················· ····················· ···················· ···················· ····················
P 9631 hCoV-19/Denmark/DCGC-6511/2020|EPI_ISL_619411|2020 ···················· ···················· ····················· ···················· ···················· ····················
P 9632 hCoV-19/Denmark/DCGC-6512/2020|EPI_ISL_619412|2020 ···················· ···················· ····················· ···················· ···················· ····················
P 9633 hCoV-19/Denmark/DCGC-6513/2020|EPI_ISL_619413|2020 ···················· ···················· ····················· ···················· ···················· ····················
P 9634 hCoV-19/Denmark/DCGC-6514/2020|EPI_ISL_619414|2020 ···················· ···················· ····················· ···················· ···················· ····················
P 9635 hCoV-19/Denmark/DCGC-6516/2020|EPI_ISL_619416|2020 ···················· ···················· ····················· ···················· ···················· ····················
P 9636 hCoV-19/Denmark/DCGC-6520/2020|EPI_ISL_619420|2020 ···················· ···················· ····················· ···················· ···················· ····················

| | | | | | | | | |
|---|---|---|---|---|---|---|---|---|
| P | 9637 | hCoV-19/Denmark/DCGC-6522/2020\|EPI_ISL_619422\|2020 | ···················· | ···················· | ····················· | ···················· | ···················· | ···················· |
| P | 9638 | hCoV-19/Denmark/DCGC-6523/2020\|EPI_ISL_619423\|2020 | ···················· | ···················· | ····················· | ···················· | ···················· | ···················· |
| P | 9639 | hCoV-19/Denmark/DCGC-6525/2020\|EPI_ISL_619425\|2020 | ···················· | ···················· | ····················· | ···················· | ···················· | ···················· |
| P | 9640 | hCoV-19/Denmark/DCGC-6526/2020\|EPI_ISL_619426\|2020 | ···················· | ···················· | ····················· | ···················· | ···················· | ···················· |
| P | 9641 | hCoV-19/Denmark/DCGC-6527/2020\|EPI_ISL_619427\|2020 | ···················· | ···················· | ····················· | ···················· | ···················· | ···················· |
| P | 9642 | hCoV-19/Denmark/DCGC-6528/2020\|EPI_ISL_619428\|2020 | ···················· | ···················· | ····················· | ···················· | ···················· | ···················· |
| P | 9643 | hCoV-19/Denmark/DCGC-6529/2020\|EPI_ISL_619429\|2020 | ···················· | ···················· | ····················· | ···················· | ···················· | ···················· |
| P | 9644 | hCoV-19/Denmark/DCGC-6530/2020\|EPI_ISL_619430\|2020 | ···················· | ···················· | ····················· | ···················· | ···T················ | ···················· |
| P | 9645 | hCoV-19/Denmark/DCGC-6531/2020\|EPI_ISL_619431\|2020 | ···················· | ···················· | ····················· | ···················· | ···················· | ···················· |
| P | 9646 | hCoV-19/Denmark/DCGC-6532/2020\|EPI_ISL_619432\|2020 | ···················· | ···················· | ····················· | ···················· | ···················· | ···················· |
| P | 9647 | hCoV-19/Denmark/DCGC-6533/2020\|EPI_ISL_619433\|2020 | ···················· | ·A·················· | ····················· | ···················· | ···················· | ···················· |
| P | 9648 | hCoV-19/Denmark/DCGC-6534/2020\|EPI_ISL_619434\|2020 | ···················· | ···················· | ····················· | ···················· | ···················· | ···················· |
| P | 9649 | hCoV-19/Denmark/DCGC-6535/2020\|EPI_ISL_619435\|2020 | ···················· | ···················· | ····················· | ···················· | ···················· | ···················· |
| P | 9650 | hCoV-19/Denmark/DCGC-6536/2020\|EPI_ISL_619436\|2020 | ···················· | ···················· | ····················· | ···················· | ···················· | ···················· |
| P | 9651 | hCoV-19/Denmark/DCGC-6537/2020\|EPI_ISL_619437\|2020 | ···················· | ···················· | ····················· | ···················· | ···················· | ···················· |
| P | 9652 | hCoV-19/Denmark/DCGC-6538/2020\|EPI_ISL_619438\|2020 | ···················· | ···················· | ····················· | ···················· | ···················· | ···················· |
| P | 9653 | hCoV-19/Denmark/DCGC-6540/2020\|EPI_ISL_619440\|2020 | ···················· | ···················· | ····················· | ···················· | ···················· | ···················· |
| P | 9654 | hCoV-19/Denmark/DCGC-6542/2020\|EPI_ISL_619442\|2020 | ···················· | ···················· | ····················· | ···················· | ···················· | ···················· |
| P | 9655 | hCoV-19/Denmark/DCGC-6544/2020\|EPI_ISL_619444\|2020 | ···················· | ···················· | ····················· | ···················· | ···················· | ···················· |
| P | 9656 | hCoV-19/Denmark/DCGC-6545/2020\|EPI_ISL_619445\|2020 | ···················· | ···················· | ····················· | ···················· | ···················· | ···················· |
| P | 9657 | hCoV-19/Denmark/DCGC-6547/2020\|EPI_ISL_619447\|2020 | ···················· | ···················· | ····················· | ···················· | ···················· | ···················· |
| P | 9658 | hCoV-19/Denmark/DCGC-6775/2020\|EPI_ISL_619471\|2020 | ···················· | ···················· | ····················· | ···················· | ···················· | ···················· |
| P | 9659 | hCoV-19/Denmark/DCGC-6776/2020\|EPI_ISL_619472\|2020 | ···················· | ···················· | ····················· | ···················· | ···················· | ···················· |
| P | 9660 | hCoV-19/Denmark/DCGC-6779/2020\|EPI_ISL_619475\|2020 | ···················· | ···················· | ····················· | ···················· | ···················· | ···················· |
| P | 9661 | hCoV-19/Denmark/DCGC-6780/2020\|EPI_ISL_619476\|2020 | ···················· | ···················· | ····················· | ···················· | ···················· | ···················· |
| P | 9662 | hCoV-19/Denmark/DCGC-6788/2020\|EPI_ISL_619484\|2020 | ···················· | ···················· | ····················· | ···················· | ···················· | ···················· |
| P | 9663 | hCoV-19/Denmark/DCGC-6789/2020\|EPI_ISL_619485\|2020 | ···················· | ···················· | ····················· | ···················· | ···················· | ···················· |
| P | 9664 | hCoV-19/Denmark/DCGC-6790/2020\|EPI_ISL_619486\|2020 | ···················· | ···················· | ····················· | ···················· | ···················· | ···················· |
| P | 9665 | hCoV-19/Denmark/DCGC-6793/2020\|EPI_ISL_619489\|2020 | ···················· | ···················· | ····················· | ···················· | ···················· | ···················· |
| P | 9666 | hCoV-19/Denmark/DCGC-6795/2020\|EPI_ISL_619491\|2020 | ···················· | ···················· | ····················· | ···················· | ···················· | ···················· |
| P | 9667 | hCoV-19/Denmark/DCGC-1286/2020\|EPI_ISL_619492\|2020 | ···················· | ···················· | ····················· | ···················· | ···················· | ···················· |
| P | 9668 | hCoV-19/Denmark/DCGC-6796/2020\|EPI_ISL_619493\|2020 | ···················· | ···················· | ····················· | ···················· | ···················· | ···················· |
| P | 9669 | hCoV-19/Denmark/DCGC-6797/2020\|EPI_ISL_619494\|2020 | ···················· | ···················· | ····················· | ···················· | ···················· | ···················· |
| P | 9670 | hCoV-19/Denmark/DCGC-6798/2020\|EPI_ISL_619495\|2020 | ···················· | ·A·················· | ····················· | ···················· | ···················· | ··A················· |
| P | 9671 | hCoV-19/Denmark/DCGC-6799/2020\|EPI_ISL_619496\|2020 | ···················· | ···················· | ····················· | ···················· | ···················· | ···················· |
| P | 9672 | hCoV-19/Denmark/DCGC-6800/2020\|EPI_ISL_619497\|2020 | ···················· | ···················· | ····················· | ···················· | ···················· | ···················· |
| P | 9673 | hCoV-19/Denmark/DCGC-6801/2020\|EPI_ISL_619498\|2020 | ···················· | ···················· | ····················· | ···················· | ···················· | ···················· |
| P | 9674 | hCoV-19/Denmark/DCGC-6802/2020\|EPI_ISL_619499\|2020 | ···················· | ···················· | ····················· | ···················· | ···················· | ···················· |
| P | 9675 | hCoV-19/Denmark/DCGC-6500/2020\|EPI_ISL_620400\|2020 | ···················· | ···················· | ····················· | ···················· | ···················· | ···················· |
| P | 9676 | hCoV-19/Denmark/DCGC-6501/2020\|EPI_ISL_620401\|2020 | ···················· | ···················· | ····················· | ···················· | ···················· | ···················· |
| P | 9677 | hCoV-19/Denmark/DCGC-6503/2020\|EPI_ISL_620403\|2020 | ···················· | ···················· | ····················· | ···················· | ···················· | ···················· |
| P | 9678 | hCoV-19/Denmark/DCGC-6504/2020\|EPI_ISL_620404\|2020 | ···················· | ···················· | ····················· | ···················· | ···················· | ···················· |
| P | 9679 | hCoV-19/Denmark/DCGC-5184/2020\|EPI_ISL_621722\|2020 | ···················· | ···················· | ····················· | ···················· | ···················· | ···················· |
| P | 9680 | hCoV-19/Denmark/DCGC-5186/2020\|EPI_ISL_621724\|2020 | ···················· | ···················· | ····················· | ···················· | ···················· | ···················· |
| P | 9681 | hCoV-19/Denmark/DCGC-5187/2020\|EPI_ISL_621725\|2020 | ···················· | ···················· | ····················· | ···················· | ···················· | ···················· |
| P | 9682 | hCoV-19/Denmark/DCGC-5188/2020\|EPI_ISL_621726\|2020 | ···················· | ···················· | ····················· | ···················· | ···················· | ···················· |
| P | 9683 | hCoV-19/Denmark/DCGC-5189/2020\|EPI_ISL_621727\|2020 | ···················· | ···················· | ····················· | ···················· | ···················· | ···················· |
| P | 9684 | hCoV-19/Denmark/DCGC-5190/2020\|EPI_ISL_621728\|2020 | ···················· | ···················· | ····················· | ···················· | ···················· | ···················· |
| P | 9685 | hCoV-19/Denmark/DCGC-5191/2020\|EPI_ISL_621729\|2020 | ···················· | ···················· | ····················· | ···················· | ···················· | ···················· |
| P | 9686 | hCoV-19/Denmark/DCGC-5192/2020\|EPI_ISL_621730\|2020 | ···················· | ···················· | ····················· | ···················· | ···················· | ···················· |
| P | 9687 | hCoV-19/Denmark/DCGC-5193/2020\|EPI_ISL_621731\|2020 | ···················· | ···················· | ····················· | ···················· | ···················· | ···················· |
| P | 9688 | hCoV-19/Denmark/DCGC-5194/2020\|EPI_ISL_621732\|2020 | ···················· | ···················· | ····················· | ···················· | ···················· | ···················· |
| P | 9689 | hCoV-19/Denmark/DCGC-5195/2020\|EPI_ISL_621733\|2020 | ···················· | ···················· | ····················· | ···················· | ···················· | ···················· |
| P | 9690 | hCoV-19/Denmark/DCGC-5196/2020\|EPI_ISL_621734\|2020 | ···················· | ···················· | ····················· | ···················· | ···················· | ···················· |
| P | 9691 | hCoV-19/Denmark/DCGC-5197/2020\|EPI_ISL_621735\|2020 | ···················· | ···················· | ····················· | ···················· | ···················· | ···················· |
| P | 9692 | hCoV-19/Denmark/DCGC-5199/2020\|EPI_ISL_621737\|2020 | ···················· | ···················· | ····················· | ···················· | ···················· | ···················· |
| P | 9693 | hCoV-19/Denmark/DCGC-5200/2020\|EPI_ISL_621738\|2020 | ···················· | ···················· | ····················· | ···················· | ···················· | ···················· |
| P | 9694 | hCoV-19/Denmark/DCGC-5201/2020\|EPI_ISL_621739\|2020 | ···················· | ···················· | ····················· | ···················· | ···················· | ···················· |
| P | 9695 | hCoV-19/Denmark/DCGC-5202/2020\|EPI_ISL_621740\|2020 | ···················· | ···················· | ····················· | ···················· | ···················· | ···················· |
| P | 9696 | hCoV-19/Denmark/DCGC-5203/2020\|EPI_ISL_621741\|2020 | ···················· | ···················· | ····················· | ···················· | ···················· | ···················· |
| P | 9697 | hCoV-19/Denmark/DCGC-5204/2020\|EPI_ISL_621742\|2020 | ···················· | ···················· | ····················· | ···················· | ···················· | ···················· |
| P | 9698 | hCoV-19/Denmark/DCGC-5205/2020\|EPI_ISL_621743\|2020 | ···················· | ···················· | ····················· | ···················· | ···················· | ···················· |
| P | 9699 | hCoV-19/Denmark/DCGC-5206/2020\|EPI_ISL_621744\|2020 | ···················· | ···················· | ····················· | ···················· | ···················· | ···················· |
| P | 9700 | hCoV-19/Denmark/DCGC-5209/2020\|EPI_ISL_621747\|2020 | ···················· | ···················· | ····················· | ···················· | ···················· | ···················· |
| P | 9701 | hCoV-19/Denmark/DCGC-5210/2020\|EPI_ISL_621748\|2020 | ···················· | ···················· | ····················· | ···················· | ···················· | ···················· |
| P | 9702 | hCoV-19/Denmark/DCGC-5211/2020\|EPI_ISL_621749\|2020 | ···················· | ···················· | ····················· | ···················· | ···················· | ···················· |
| P | 9703 | hCoV-19/Denmark/DCGC-5212/2020\|EPI_ISL_621750\|2020 | ···················· | ···················· | ····················· | ···················· | ···················· | ···················· |
| P | 9704 | hCoV-19/Denmark/DCGC-5213/2020\|EPI_ISL_621751\|2020 | ···················· | ·A·················· | ····················· | ···················· | ···················· | ···················· |
| P | 9705 | hCoV-19/Denmark/DCGC-5214/2020\|EPI_ISL_621752\|2020 | ···················· | ·A·················· | ····················· | ···················· | ···················· | ···················· |
| P | 9706 | hCoV-19/Denmark/DCGC-5215/2020\|EPI_ISL_621753\|2020 | ···················· | ···················· | ····················· | ···················· | ···················· | ···················· |
| P | 9707 | hCoV-19/Denmark/DCGC-5217/2020\|EPI_ISL_621755\|2020 | ···················· | ···················· | ····················· | ···················· | ···················· | ···················· |
| P | 9708 | hCoV-19/Denmark/DCGC-5218/2020\|EPI_ISL_621756\|2020 | ···················· | ···················· | ····················· | ···················· | ···················· | ···················· |
| P | 9709 | hCoV-19/Denmark/DCGC-5219/2020\|EPI_ISL_621757\|2020 | ···················· | ···················· | ····················· | ···················· | ···················· | ···················· |
| P | 9710 | hCoV-19/Denmark/DCGC-5220/2020\|EPI_ISL_621758\|2020 | ···················· | ···················· | ····················· | ···················· | ···················· | ···················· |
| P | 9711 | hCoV-19/Denmark/DCGC-5221/2020\|EPI_ISL_621759\|2020 | ···················· | ···················· | ····················· | ···················· | ···················· | ···················· |
| P | 9712 | hCoV-19/Denmark/DCGC-5222/2020\|EPI_ISL_621760\|2020 | ···················· | ···················· | ····················· | ···················· | ···················· | ···················· |
| P | 9713 | hCoV-19/Denmark/DCGC-5223/2020\|EPI_ISL_621761\|2020 | ···················· | ···················· | ····················· | ···················· | ···················· | ···················· |
| P | 9714 | hCoV-19/Denmark/DCGC-5009/2020\|EPI_ISL_621762\|2020 | ···················· | ···················· | ····················· | ···················· | ···················· | ···················· |
| P | 9715 | hCoV-19/Denmark/DCGC-5022/2020\|EPI_ISL_621763\|2020 | ···················· | ···················· | ····················· | ···················· | ···················· | ···················· |
| P | 9716 | hCoV-19/Denmark/DCGC-5029/2020\|EPI_ISL_621764\|2020 | ···················· | ···················· | ····················· | ···················· | ···················· | ···················· |

| | | | | | | | | |
|---|---|---|---|---|---|---|---|---|
| P | 9717 | hCoV-19/Denmark/DCGC-5035/2020\|EPI_ISL_621765\|2020 | .................... | .................... | ..................... | .................... | .................... | .................... |
| P | 9718 | hCoV-19/Denmark/DCGC-4987/2020\|EPI_ISL_621788\|2020 | .................... | .................... | ..................... | .................... | .................... | .................... |
| P | 9719 | hCoV-19/Denmark/DCGC-4995/2020\|EPI_ISL_621789\|2020 | .................... | .................... | ..................... | .................... | .................... | .................... |
| P | 9720 | hCoV-19/Denmark/DCGC-5006/2020\|EPI_ISL_621790\|2020 | .................... | .................... | ..................... | .................... | .................... | .................... |
| P | 9721 | hCoV-19/Denmark/DCGC-5011/2020\|EPI_ISL_621791\|2020 | .................... | .................... | ..................... | .................... | .................... | .................... |
| P | 9722 | hCoV-19/Denmark/DCGC-5018/2020\|EPI_ISL_621792\|2020 | .................... | .................... | ..................... | .................... | .................... | .................... |
| P | 9723 | hCoV-19/Denmark/DCGC-5025/2020\|EPI_ISL_621793\|2020 | .................... | .................... | ..................... | .................... | .................... | .................... |
| P | 9724 | hCoV-19/Denmark/DCGC-5031/2020\|EPI_ISL_621794\|2020 | .................... | .................... | ..................... | .................... | .................... | .................... |
| P | 9725 | hCoV-19/Denmark/DCGC-5036/2020\|EPI_ISL_621795\|2020 | .................... | .................... | ..................... | .................... | .................... | .................... |
| P | 9726 | hCoV-19/Denmark/DCGC-5043/2020\|EPI_ISL_621796\|2020 | .................... | .................... | ..................... | .................... | .................... | .................... |
| P | 9727 | hCoV-19/Denmark/DCGC-5049/2020\|EPI_ISL_621797\|2020 | .................... | .................... | ..................... | .................... | .................... | .................... |
| P | 9728 | hCoV-19/Denmark/DCGC-4988/2020\|EPI_ISL_621798\|2020 | .................... | .................... | ..................... | .................... | .................... | .................... |
| P | 9729 | hCoV-19/Denmark/DCGC-3098/2020\|EPI_ISL_615914\|2020 | .................... | .................... | ..................... | .................... | .................... | .................... |
| P | 9730 | hCoV-19/Denmark/DCGC-3101/2020\|EPI_ISL_615917\|2020 | .................... | .................... | ..................... | .................... | .................... | .................... |
| P | 9731 | hCoV-19/Denmark/DCGC-3102/2020\|EPI_ISL_615918\|2020 | .................... | .................... | ..................... | .................... | .................... | .................... |
| P | 9732 | hCoV-19/Denmark/DCGC-3103/2020\|EPI_ISL_615919\|2020 | .................... | .................... | ..................... | .................... | .................... | .................... |
| P | 9733 | hCoV-19/Denmark/DCGC-1773/2020\|EPI_ISL_618192\|2020 | .................... | .................... | ..................... | .................... | .................... | .................... |

Group 5- Oct 2020-Dec 2020

| A | B | C | D | E | F | G | H | I | J | K | L |
|---|---|---|---|---|---|---|---|---|---|---|---|
| Database | Database ID | GISAID ID | GCTGCAATCGTGCTACAACT | TGAACTGTTGCGACTACGTG | TTCGGAAGAGACAGGTACGTT | CACACAATCGAAGCGCAGTA | GCTGGTGCTGCAGCTTATTA | AGGGTCAAGTGCACAGTCTA | mismatches? | mismatches? | primer set? |
| Q | 1 | hCoV-19/England/PORT-2D5BA7/2020\|EPI_ISL_628320\|20 | ···················· | ···················· | ···················· | ···················· | ·····T·············· | ···················· | | | |
| Q | 2 | hCoV-19/England/PORT-2D5D47/2020\|EPI_ISL_628321\|20 | ···················· | ···················· | ···················· | ···················· | ·····T·············· | ···················· | | | |
| Q | 3 | hCoV-19/England/PORT-2D5DDE/2020\|EPI_ISL_628322\|20 | ···················· | ···················· | ···················· | ···················· | ·····T·············· | ···················· | | | |
| Q | 4 | hCoV-19/England/PORT-2D7158/2020\|EPI_ISL_628323\|20 | ···················· | ···················· | ···················· | ···················· | ·····T·············· | ···················· | | | |
| Q | 5 | hCoV-19/England/PORT-2D7167/2020\|EPI_ISL_628324\|20 | ···················· | ···················· | ···················· | ···················· | ·····T·············· | ···················· | | | |
| Q | 6 | hCoV-19/England/PORT-2D7185/2020\|EPI_ISL_628325\|20 | ···················· | ···················· | ···················· | ···················· | ···················· | ···················· | | | |
| Q | 7 | hCoV-19/England/PORT-2D7194/2020\|EPI_ISL_628326\|20 | ···················· | ···················· | ···················· | ···················· | ···················· | ···················· | | | |
| Q | 8 | hCoV-19/England/PORT-2D71B2/2020\|EPI_ISL_628327\|20 | ···················· | ···················· | ···················· | ···················· | ···················· | ···················· | | | |
| Q | 10 | hCoV-19/England/PORT-2D7282/2020\|EPI_ISL_628329\|20 | ···················· | ···················· | ···················· | ···················· | ···················· | ···················· | | | |
| Q | 11 | hCoV-19/England/PORT-2D72EC/2020\|EPI_ISL_628330\|20 | ···················· | ···················· | ···················· | ···················· | ·····T·············· | ···················· | | | |
| Q | 12 | hCoV-19/England/PORT-2D7307/2020\|EPI_ISL_628331\|20 | ···················· | ···················· | ···················· | ···················· | ···················· | ···················· | | | |
| Q | 13 | hCoV-19/England/PORT-2D7361/2020\|EPI_ISL_628332\|20 | ···················· | ···················· | ···················· | ···················· | ·····T·············· | ···················· | | | |
| Q | 14 | hCoV-19/England/PORT-2D7370/2020\|EPI_ISL_628333\|20 | ···················· | ···················· | ···················· | ···················· | ···················· | ···················· | | | |
| Q | 15 | hCoV-19/Scotland/QEUH-A321F9/2020\|EPI_ISL_628334\|2 | ···················· | ···················· | ···················· | ···················· | ···················· | ···················· | | | |
| Q | 16 | hCoV-19/Scotland/QEUH-A32214/2020\|EPI_ISL_628335\|2 | ···················· | ···················· | ···················· | ···················· | ···················· | ···················· | | | |
| Q | 17 | hCoV-19/England/QEUH-A32223/2020\|EPI_ISL_628336\|20 | ···················· | ·A·················· | ···················· | ···················· | ···················· | ···················· | | | |
| Q | 18 | hCoV-19/Scotland/QEUH-A32232/2020\|EPI_ISL_628337\|2 | ···················· | ···················· | ···················· | ···················· | ···················· | ···················· | | | |
| Q | 19 | hCoV-19/England/QEUH-A3228D/2020\|EPI_ISL_628338\|20 | ···················· | ···················· | ···················· | ···················· | ···················· | ···················· | | | |
| Q | 20 | hCoV-19/Scotland/QEUH-A3229C/2020\|EPI_ISL_628339\|2 | ···················· | ···················· | ···················· | ···················· | ···················· | ···················· | | | |
| Q | 21 | hCoV-19/Scotland/QEUH-A3233F/2020\|EPI_ISL_628347\|2 | ···················· | ···················· | ···················· | ···················· | ···················· | ···················· | | | |
| Q | 22 | hCoV-19/England/QEUH-A3236C/2020\|EPI_ISL_628348\|20 | ···················· | ···················· | ···················· | ···················· | ···················· | ···················· | | | |
| Q | 23 | hCoV-19/England/QEUH-A3238A/2020\|EPI_ISL_628349\|20 | ···················· | ···················· | ···················· | ···················· | ···················· | ···················· | | | |
| Q | 24 | hCoV-19/England/QEUH-A323B7/2020\|EPI_ISL_628350\|20 | ···················· | ···················· | ···················· | ···················· | ···················· | ···················· | | | |
| Q | 25 | hCoV-19/England/QEUH-A323F3/2020\|EPI_ISL_628351\|20 | ···················· | ···················· | ···················· | ···················· | ···················· | ···················· | | | |
| Q | 26 | hCoV-19/England/QEUH-A3240F/2020\|EPI_ISL_628352\|20 | ···················· | ·A·················· | ···················· | ···················· | ···················· | ··A················· | | | |
| Q | 27 | hCoV-19/England/QEUH-A3241E/2020\|EPI_ISL_628353\|20 | ···················· | ···················· | ···················· | ···················· | ···················· | ···················· | | | |
| Q | 28 | hCoV-19/England/QEUH-A32636/2020\|EPI_ISL_628354\|20 | ···················· | ···················· | ···················· | ···················· | ···················· | ···················· | | | |
| Q | 29 | hCoV-19/England/QEUH-A32645/2020\|EPI_ISL_628355\|20 | ···················· | ···················· | ···················· | ···················· | ·····T·············· | ···················· | | | |
| Q | 30 | hCoV-19/England/QEUH-A32663/2020\|EPI_ISL_628356\|20 | ···················· | ···················· | ···················· | ···················· | ···················· | ···················· | | | |
| Q | 31 | hCoV-19/England/QEUH-A32690/2020\|EPI_ISL_628357\|20 | ···················· | ···················· | ···················· | ···················· | ···················· | ···················· | | | |
| Q | 32 | hCoV-19/England/QEUH-A326AF/2020\|EPI_ISL_628358\|20 | ···················· | ···················· | ···················· | ···················· | ···················· | ···················· | | | |
| Q | 33 | hCoV-19/England/QEUH-A326BE/2020\|EPI_ISL_628359\|20 | ···················· | ···················· | ···················· | ···················· | ···················· | ···················· | | | |
| Q | 34 | hCoV-19/England/MILK-ACEEFB/2020\|EPI_ISL_629601\|20 | ···················· | ···················· | ···················· | ···················· | ···················· | ···················· | | | |
| Q | 35 | hCoV-19/England/MILK-ACEB12/2020\|EPI_ISL_629603\|20 | ···················· | ···················· | ···················· | ···················· | ···················· | ···················· | | | |
| Q | 36 | hCoV-19/England/MILK-ACEEBF/2020\|EPI_ISL_629604\|20 | ···················· | ···················· | ···················· | ···················· | ···················· | ···················· | | | |
| Q | 37 | hCoV-19/England/MILK-ACF15B/2020\|EPI_ISL_629605\|20 | ···················· | ···················· | ···················· | ···················· | ···················· | ···················· | | | |
| Q | 38 | hCoV-19/England/MILK-AC8DE5/2020\|EPI_ISL_629606\|20 | ···················· | ···················· | ···················· | ···················· | ···················· | ···················· | | | |
| Q | 39 | hCoV-19/England/MILK-ACEDD0/2020\|EPI_ISL_629650\|20 | ···················· | ···················· | ···················· | ···················· | ···················· | ···················· | | | |
| Q | 40 | hCoV-19/England/MILK-ACF3FB/2020\|EPI_ISL_629651\|20 | ···················· | ·A·················· | ···················· | ···················· | ···················· | ··A················· | | | |
| Q | 41 | hCoV-19/England/MILK-AC89D2/2020\|EPI_ISL_629652\|20 | ···················· | ···················· | ···················· | ···················· | ···················· | ···················· | | | |
| Q | 42 | hCoV-19/England/MILK-ACF4F8/2020\|EPI_ISL_629653\|20 | ···················· | ···················· | ···················· | ···················· | ···················· | ···················· | | | |
| Q | 43 | hCoV-19/England/MILK-AC9081/2020\|EPI_ISL_629655\|20 | ···················· | ···················· | ···················· | ···················· | ···················· | ···················· | | | |
| Q | 44 | hCoV-19/England/MILK-AC84B3/2020\|EPI_ISL_629656\|20 | ···················· | ·A·················· | ···················· | ···················· | ···················· | ··A················· | | | |
| Q | 45 | hCoV-19/England/MILK-AC8978/2020\|EPI_ISL_629657\|20 | ···················· | ···················· | ···················· | ···················· | ···················· | ···················· | | | |
| Q | 46 | hCoV-19/England/MILK-ACFA32/2020\|EPI_ISL_629658\|20 | ···················· | ···················· | ···················· | ···················· | ···················· | ···················· | | | |
| Q | 47 | hCoV-19/England/MILK-ACF4CB/2020\|EPI_ISL_629659\|20 | ···················· | ·A·················· | ···················· | ···················· | ···················· | ···················· | | | |
| Q | 48 | hCoV-19/England/MILK-ACF22B/2020\|EPI_ISL_629660\|20 | ···················· | ···················· | ···················· | ···················· | ···················· | ···················· | | | |
| Q | 49 | hCoV-19/England/MILK-ACEA51/2020\|EPI_ISL_629661\|20 | ···················· | ···················· | ···················· | ···················· | ···················· | ···················· | | | |
| Q | 50 | hCoV-19/England/MILK-AC7CAD/2020\|EPI_ISL_629664\|20 | ···················· | ···················· | ···················· | ···················· | ···················· | ···················· | | | |
| Q | 51 | hCoV-19/England/MILK-AC7F0E/2020\|EPI_ISL_629667\|20 | ···················· | ···················· | ···················· | ···················· | ···················· | ···················· | | | |
| Q | 52 | hCoV-19/England/MILK-ACEE19/2020\|EPI_ISL_629669\|20 | ···················· | ···················· | ···················· | ···················· | ···················· | ···················· | | | |
| Q | 53 | hCoV-19/England/MILK-ACEE64/2020\|EPI_ISL_629679\|20 | ···················· | ···················· | ···················· | ···················· | ···················· | ···················· | | | |
| Q | 54 | hCoV-19/England/MILK-ACFA05/2020\|EPI_ISL_629680\|20 | ···················· | ···················· | ···················· | ···················· | ···················· | ···················· | | | |
| Q | 55 | hCoV-19/England/MILK-AC932E/2020\|EPI_ISL_629681\|20 | ···················· | ···················· | ···················· | ···················· | ···················· | ···················· | | | |
| Q | 56 | hCoV-19/England/MILK-AC826E/2020\|EPI_ISL_629682\|20 | ···················· | ···················· | ···················· | ···················· | ···················· | ···················· | | | |
| Q | 57 | hCoV-19/England/MILK-AC8F76/2020\|EPI_ISL_629684\|20 | ···················· | ···················· | ···················· | ···················· | ···················· | ···················· | | | |
| Q | 58 | hCoV-19/England/MILK-ACF11F/2020\|EPI_ISL_629685\|20 | ···················· | ···················· | ···················· | ···················· | ···················· | ···················· | | | |
| Q | 59 | hCoV-19/England/MILK-ACF9EA/2020\|EPI_ISL_629686\|20 | ···················· | ···················· | ···················· | ···················· | ···················· | ···················· | | | |
| Q | 60 | hCoV-19/England/MILK-AC91D9/2020\|EPI_ISL_629687\|20 | ···················· | ···················· | ···················· | ···················· | ···················· | ···················· | | | |
| Q | 61 | hCoV-19/England/MILK-ACE848/2020\|EPI_ISL_629689\|20 | ···················· | ···················· | ···················· | ···················· | ···················· | ···················· | | | |
| Q | 62 | hCoV-19/England/ALDP-A68646/2020\|EPI_ISL_630633\|20 | ···················· | ···················· | ···················· | ···················· | ···················· | ···················· | | | |
| Q | 63 | hCoV-19/England/ALDP-A6795D/2020\|EPI_ISL_630635\|20 | ···················· | ···················· | ···················· | ···················· | ···················· | ···················· | | | |
| Q | 64 | hCoV-19/England/ALDP-A68628/2020\|EPI_ISL_630636\|20 | ···················· | ···················· | ···················· | ···················· | ···················· | ···················· | | | |
| Q | 65 | hCoV-19/England/ALDP-A67920/2020\|EPI_ISL_630637\|20 | ···················· | ···················· | ···················· | ···················· | ···················· | ···················· | | | |
| Q | 66 | hCoV-19/England/ALDP-A670EF/2020\|EPI_ISL_630638\|20 | ···················· | ···················· | ···················· | ···················· | ···················· | ···················· | | | |
| Q | 67 | hCoV-19/England/ALDP-A67744/2020\|EPI_ISL_630639\|20 | ···················· | ···················· | ···················· | ···················· | ···················· | ···················· | | | |
| Q | 68 | hCoV-19/England/ALDP-A68619/2020\|EPI_ISL_630640\|20 | ···················· | ···················· | ···················· | ···················· | ···················· | ···················· | | | |
| Q | 69 | hCoV-19/England/ALDP-A66E40/2020\|EPI_ISL_630641\|20 | ···················· | ·A·················· | ···················· | ···················· | ···················· | ··A················· | | | |
| Q | 70 | hCoV-19/England/ALDP-A671BF/2020\|EPI_ISL_630642\|20 | ···················· | ···················· | ···················· | ···················· | ···················· | ···················· | | | |
| Q | 71 | hCoV-19/England/ALDP-A6788D/2020\|EPI_ISL_630643\|20 | ···················· | ···················· | ···················· | ···················· | ···················· | ···················· | | | |

| | | | | | | | | |
|---|---|---|---|---|---|---|---|---|
| Q | 72 | hCoV-19/England/ALDP-A67841/2020\|EPI_ISL_630644\|20 | ···················· | ···················· | ···················· | ···················· | ···················· | ···················· |
| Q | 73 | hCoV-19/England/ALDP-A66824/2020\|EPI_ISL_630645\|20 | ···················· | ·A·················· | ···················· | ···················· | ···················· | ··A················· |
| Q | 74 | hCoV-19/England/ALDP-A6761A/2020\|EPI_ISL_630646\|20 | ···················· | ···················· | ···················· | ···················· | ···················· | ···················· |
| Q | 75 | hCoV-19/England/ALDP-A67638/2020\|EPI_ISL_630647\|20 | ···················· | ···················· | ···················· | ···················· | ···················· | ···················· |
| Q | 76 | hCoV-19/England/ALDP-A678D8/2020\|EPI_ISL_630648\|20 | ···················· | ···················· | ···················· | ···················· | ···················· | ···················· |
| Q | 77 | hCoV-19/England/ALDP-A66A97/2020\|EPI_ISL_630649\|20 | ···················· | ···················· | ···················· | ···················· | ···················· | ···················· |
| Q | 78 | hCoV-19/England/ALDP-A67C90/2020\|EPI_ISL_630651\|20 | ···················· | ···················· | ···················· | ···················· | ···················· | ···················· |
| Q | 79 | hCoV-19/England/ALDP-A67CFA/2020\|EPI_ISL_630652\|20 | ···················· | ·A·················· | ···················· | ···················· | ···················· | ··A················· |
| Q | 80 | hCoV-19/England/ALDP-A66C82/2020\|EPI_ISL_630653\|20 | ···················· | ···················· | ···················· | ···················· | ···················· | ···················· |
| Q | 81 | hCoV-19/England/ALDP-A6744D/2020\|EPI_ISL_630658\|20 | ···················· | ···················· | ···················· | ···················· | ···················· | ···················· |
| Q | 82 | hCoV-19/England/ALDP-A67322/2020\|EPI_ISL_630659\|20 | ···················· | ···················· | ···················· | ···················· | ···················· | ···················· |
| Q | 83 | hCoV-19/England/ALDP-A66A2E/2020\|EPI_ISL_630660\|20 | ···················· | ···················· | ···················· | ···················· | ···················· | ···················· |
| Q | 84 | hCoV-19/England/ALDP-A670FE/2020\|EPI_ISL_630661\|20 | ···················· | ···················· | ···················· | ···················· | ···················· | ···················· |
| Q | 85 | hCoV-19/England/ALDP-A674A7/2020\|EPI_ISL_630662\|20 | ···················· | ···················· | ···················· | ···················· | ···················· | ···················· |
| Q | 86 | hCoV-19/England/ALDP-A6760B/2020\|EPI_ISL_630663\|20 | ···················· | ···················· | ···················· | ···················· | ···················· | ···················· |
| Q | 87 | hCoV-19/England/ALDP-A66F2F/2020\|EPI_ISL_630664\|20 | ···················· | ···················· | ···················· | ···················· | ···················· | ···················· |
| Q | 88 | hCoV-19/England/ALDP-A67A1E/2020\|EPI_ISL_630665\|20 | ···················· | ···················· | ···················· | ···················· | ···················· | ···················· |
| Q | 89 | hCoV-19/England/ALDP-A66A1F/2020\|EPI_ISL_630667\|20 | ···················· | ···················· | ···················· | ···················· | ···················· | ···················· |
| Q | 90 | hCoV-19/England/ALDP-A67C72/2020\|EPI_ISL_630668\|20 | ···················· | ···················· | ···················· | ···················· | ···················· | ···················· |
| Q | 91 | hCoV-19/England/ALDP-A674D4/2020\|EPI_ISL_630669\|20 | ···················· | ···················· | ···················· | ···················· | ···················· | ··A················· |
| Q | 92 | hCoV-19/England/ALDP-A67128/2020\|EPI_ISL_630670\|20 | ···················· | ···················· | ···················· | ···················· | ···················· | ···················· |
| Q | 93 | hCoV-19/England/ALDP-A67C63/2020\|EPI_ISL_630671\|20 | ···················· | ···················· | ···················· | ···················· | ···················· | ···················· |
| Q | 94 | hCoV-19/England/ALDP-A66F6B/2020\|EPI_ISL_630672\|20 | ···················· | ···················· | ···················· | ···················· | ···················· | ···················· |
| Q | 95 | hCoV-19/England/ALDP-A66BB2/2020\|EPI_ISL_630673\|20 | ···················· | ···················· | ···················· | ···················· | ···················· | ···················· |
| Q | 96 | hCoV-19/England/ALDP-A672F8/2020\|EPI_ISL_630674\|20 | ···················· | ···················· | ···················· | ···················· | ···················· | ···················· |
| Q | 97 | hCoV-19/USA/NY-NYCPHL-000419/2020\|EPI_ISL_631902\|2 | ···················· | ···················· | ···················· | ···················· | ···················· | ···················· |
| Q | 98 | hCoV-19/USA/NY-NYCPHL-000420/2020\|EPI_ISL_631903\|2 | ···················· | ···················· | ···················· | ···················· | ···················· | ···················· |
| Q | 99 | hCoV-19/USA/NY-NYCPHL-000421/2020\|EPI_ISL_631904\|2 | ···················· | ···················· | ···················· | ···················· | ···················· | ···················· |
| Q | 100 | hCoV-19/USA/NY-NYCPHL-000422/2020\|EPI_ISL_631905\|2 | ···················· | ···················· | ···················· | ···················· | ···················· | ···················· |
| Q | 101 | hCoV-19/USA/NY-NYCPHL-000423/2020\|EPI_ISL_631906\|2 | ···················· | ···················· | ···················· | ···················· | ···················· | ···················· |
| Q | 102 | hCoV-19/USA/NY-NYCPHL-000424/2020\|EPI_ISL_631907\|2 | ···················· | ···················· | ···················· | ···················· | ···················· | ···················· |
| Q | 103 | hCoV-19/USA/NY-NYCPHL-000426/2020\|EPI_ISL_631909\|2 | ···················· | ···················· | ···················· | ···················· | ···················· | ···················· |
| Q | 104 | hCoV-19/USA/NY-NYCPHL-000434/2020\|EPI_ISL_631912\|2 | ···················· | ···················· | ···················· | ···················· | ···················· | ···················· |
| Q | 105 | hCoV-19/USA/NY-NYCPHL-000440/2020\|EPI_ISL_631914\|2 | ···················· | ···················· | ···················· | ···················· | ···················· | ···················· |
| Q | 106 | hCoV-19/USA/NY-NYCPHL-000441/2020\|EPI_ISL_631915\|2 | ···················· | ···················· | ···················· | ···················· | ···················· | ···················· |
| Q | 107 | hCoV-19/USA/NY-NYCPHL-000443/2020\|EPI_ISL_631917\|2 | ···················· | ···················· | ···················· | ···················· | ···················· | ···················· |
| Q | 108 | hCoV-19/USA/NY-NYCPHL-000444/2020\|EPI_ISL_631918\|2 | ···················· | ···················· | ···················· | ···················· | ···················· | ···················· |
| Q | 109 | hCoV-19/USA/NY-NYCPHL-000445/2020\|EPI_ISL_631919\|2 | ···················· | ···················· | ···················· | ···················· | ···················· | ···················· |
| Q | 110 | hCoV-19/USA/NY-NYCPHL-000502/2020\|EPI_ISL_631963\|2 | ···················· | ···················· | ···················· | ···················· | ···················· | ···················· |
| Q | 111 | hCoV-19/USA/NY-NYCPHL-000503/2020\|EPI_ISL_631964\|2 | ···················· | ···················· | ···················· | ···················· | ···················· | ···················· |
| Q | 112 | hCoV-19/USA/NY-NYCPHL-000504/2020\|EPI_ISL_631965\|2 | ···················· | ···················· | ···················· | ···················· | ···················· | ···················· |
| Q | 113 | hCoV-19/USA/NY-NYCPHL-000505/2020\|EPI_ISL_631966\|2 | ···················· | ···················· | ···················· | ···················· | ···················· | ···················· |
| Q | 114 | hCoV-19/USA/NY-NYCPHL-000506/2020\|EPI_ISL_631967\|2 | ···················· | ···················· | ···················· | ···················· | ···················· | ···················· |
| Q | 115 | hCoV-19/USA/NY-NYCPHL-000507/2020\|EPI_ISL_631968\|2 | ···················· | ···················· | ···················· | ···················· | ···················· | ···················· |
| Q | 116 | hCoV-19/USA/NY-NYCPHL-000508/2020\|EPI_ISL_631969\|2 | ···················· | ···················· | ···················· | ···················· | ···················· | ···················· |
| Q | 117 | hCoV-19/USA/NY-NYCPHL-000509/2020\|EPI_ISL_631970\|2 | ···················· | ···················· | ···················· | ···················· | ···················· | ···················· |
| Q | 118 | hCoV-19/USA/NY-NYCPHL-000510/2020\|EPI_ISL_631971\|2 | ···················· | ···················· | ···················· | ···················· | ···················· | ···················· |
| Q | 119 | hCoV-19/USA/NY-NYCPHL-000511/2020\|EPI_ISL_631972\|2 | ···················· | ···················· | ···················· | ···················· | ···················· | ···················· |
| Q | 120 | hCoV-19/USA/NY-NYCPHL-000512/2020\|EPI_ISL_631973\|2 | ···················· | ···················· | ···················· | ···················· | ···················· | ···················· |
| Q | 121 | hCoV-19/USA/NY-NYCPHL-000513/2020\|EPI_ISL_631974\|2 | ···················· | ···················· | ···················· | ···················· | ···················· | ···················· |
| Q | 122 | hCoV-19/USA/NY-NYCPHL-000514/2020\|EPI_ISL_631975\|2 | ···················· | ···················· | ···················· | ···················· | ···················· | ···················· |
| Q | 123 | hCoV-19/USA/NY-NYCPHL-000515/2020\|EPI_ISL_631976\|2 | ···················· | ···················· | ···················· | ···················· | ···················· | ··A················· |
| Q | 124 | hCoV-19/USA/NY-NYCPHL-000516/2020\|EPI_ISL_631977\|2 | ···················· | ···················· | ···················· | ···················· | ···················· | ···················· |
| Q | 125 | hCoV-19/USA/NY-NYCPHL-000517/2020\|EPI_ISL_631978\|2 | ···················· | ···················· | ···················· | ···················· | ···················· | ···················· |
| Q | 126 | hCoV-19/USA/NY-NYCPHL-000518/2020\|EPI_ISL_631979\|2 | ···················· | ···················· | ···················· | ···················· | ···················· | ···················· |
| Q | 127 | hCoV-19/USA/NY-NYCPHL-000519/2020\|EPI_ISL_631980\|2 | ···················· | ···················· | ···················· | ···················· | ···················· | ···················· |
| Q | 128 | hCoV-19/USA/NY-NYCPHL-000520/2020\|EPI_ISL_631981\|2 | ···················· | ···················· | ···················· | ···················· | ···················· | ···················· |
| Q | 129 | hCoV-19/USA/NY-NYCPHL-000521/2020\|EPI_ISL_631982\|2 | ···················· | ···················· | ···················· | ···················· | ···················· | ···················· |
| Q | 130 | hCoV-19/USA/NY-NYCPHL-000522/2020\|EPI_ISL_631983\|2 | ···················· | ···················· | ···················· | ···················· | ···················· | ···················· |
| Q | 131 | hCoV-19/USA/NY-NYCPHL-000523/2020\|EPI_ISL_631984\|2 | ···················· | ···················· | ···················· | ···················· | ···················· | ···················· |
| Q | 132 | hCoV-19/USA/NY-NYCPHL-000528/2020\|EPI_ISL_631989\|2 | ···················· | ···················· | ···················· | ···················· | ···················· | ···················· |
| Q | 133 | hCoV-19/USA/NY-NYCPHL-000529/2020\|EPI_ISL_631990\|2 | ···················· | ···················· | ···················· | ···················· | ···················· | ···················· |
| Q | 134 | hCoV-19/USA/NY-NYCPHL-000530/2020\|EPI_ISL_631991\|2 | ···················· | ···················· | ···················· | ···················· | ···················· | ···················· |
| Q | 135 | hCoV-19/USA/NY-NYCPHL-000531/2020\|EPI_ISL_631992\|2 | ···················· | ···················· | ···················· | ···················· | ···················· | ···················· |
| Q | 136 | hCoV-19/USA/NY-NYCPHL-000532/2020\|EPI_ISL_631993\|2 | ···················· | ···················· | ···················· | ···················· | ···················· | ···················· |
| Q | 137 | hCoV-19/USA/NY-NYCPHL-000533/2020\|EPI_ISL_631994\|2 | ···················· | ···················· | ···················· | ···················· | ···················· | ···················· |
| Q | 138 | hCoV-19/USA/NY-NYCPHL-000534/2020\|EPI_ISL_631995\|2 | ···················· | ···················· | ···················· | ···················· | ···················· | ···················· |
| Q | 139 | hCoV-19/USA/NY-NYCPHL-000537/2020\|EPI_ISL_631997\|2 | ···················· | ···················· | ···················· | ···················· | ···················· | ···················· |
| Q | 140 | hCoV-19/USA/NY-NYCPHL-000538/2020\|EPI_ISL_631998\|2 | ···················· | ···················· | ···················· | ···················· | ···················· | ···················· |
| Q | 141 | hCoV-19/USA/NY-NYCPHL-000539/2020\|EPI_ISL_631999\|2 | ···················· | ···················· | ···················· | ···················· | ···················· | ···················· |
| Q | 142 | hCoV-19/England/QEUH-AD2C27/2020\|EPI_ISL_634119\|20 | ···················· | ···················· | ···················· | ···················· | ···················· | ···················· |
| Q | 143 | hCoV-19/England/QEUH-AD2CFA/2020\|EPI_ISL_634120\|20 | ···················· | ···················· | ···················· | ···················· | ···················· | ···················· |
| Q | 144 | hCoV-19/England/QEUH-AD0D17/2020\|EPI_ISL_634121\|20 | ···················· | ···················· | ···················· | ···················· | ···················· | ···················· |
| Q | 145 | hCoV-19/England/QEUH-AD3B47/2020\|EPI_ISL_634122\|20 | ···················· | ···················· | ···················· | ···················· | ···················· | ···················· |
| Q | 146 | hCoV-19/England/QEUH-AD3716/2020\|EPI_ISL_634124\|20 | ···················· | ···················· | ···················· | ···················· | ···················· | ···················· |
| Q | 147 | hCoV-19/England/QEUH-AD3048/2020\|EPI_ISL_634125\|20 | ···················· | ···················· | ···················· | ···················· | ···················· | ···················· |
| Q | 148 | hCoV-19/England/QEUH-AD302A/2020\|EPI_ISL_634126\|20 | ···················· | ···················· | ···················· | ···················· | ···················· | ···················· |
| Q | 149 | hCoV-19/England/QEUH-AD0D53/2020\|EPI_ISL_634128\|20 | ···················· | ···················· | ···················· | ···················· | ······T············· | ···················· |
| Q | 150 | hCoV-19/England/QEUH-AD3C35/2020\|EPI_ISL_634129\|20 | ···················· | ···················· | ···················· | ···················· | ···················· | ···················· |
| Q | 151 | hCoV-19/England/QEUH-AD294E/2020\|EPI_ISL_634130\|20 | ···················· | ···················· | ···················· | ···················· | ···················· | ···················· |

| | | | | | | | |
|---|---|---|---|---|---|---|---|
| Q | 152 hCoV-19/England/QEUH-AD1FE3/2020\|EPI_ISL_634131\|20 | ···················· | ···················· | ···················· | ···················· | ···················· | ···················· |
| Q | 153 hCoV-19/England/QEUH-AD5170/2020\|EPI_ISL_634132\|20 | ···················· | ···················· | ···················· | ···················· | ···················· | ···················· |
| Q | 154 hCoV-19/England/QEUH-AD50CE/2020\|EPI_ISL_634133\|20 | ···················· | ···················· | ···················· | ···················· | ···················· | ···················· |
| Q | 155 hCoV-19/England/QEUH-AD0C74/2020\|EPI_ISL_634134\|20 | ···················· | ···················· | ···················· | ···················· | ···················· | ···················· |
| Q | 156 hCoV-19/England/QEUH-AD36DD/2020\|EPI_ISL_634135\|20 | ···················· | ···················· | ···················· | ···················· | ···················· | ···················· |
| Q | 157 hCoV-19/England/QEUH-AD2C63/2020\|EPI_ISL_634136\|20 | ···················· | ···················· | ···················· | ···················· | ···················· | ···················· |
| Q | 158 hCoV-19/England/QEUH-AD29E4/2020\|EPI_ISL_634137\|20 | ···················· | ···················· | ···················· | ···················· | ···················· | ···················· |
| Q | 159 hCoV-19/England/QEUH-AD0C1A/2020\|EPI_ISL_634138\|20 | ···················· | ···················· | ···················· | ···················· | ···················· | ···················· |
| Q | 160 hCoV-19/England/QEUH-AD45DF/2020\|EPI_ISL_634139\|20 | ···················· | ···················· | ···················· | ···················· | ···················· | ···················· |
| Q | 161 hCoV-19/England/QEUH-AD29D5/2020\|EPI_ISL_634141\|20 | ···················· | ···················· | ···················· | ···················· | ···················· | ···················· |
| Q | 162 hCoV-19/England/QEUH-AD3C71/2020\|EPI_ISL_634142\|20 | ···················· | ···················· | ···················· | ···················· | ···················· | ···················· |
| Q | 163 hCoV-19/England/QEUH-AD047C/2020\|EPI_ISL_634143\|20 | ···················· | ···················· | ···················· | ···················· | ···················· | ···················· |
| Q | 164 hCoV-19/England/QEUH-AD528C/2020\|EPI_ISL_634144\|20 | ···················· | ···················· | ···················· | ···················· | ···················· | ···················· |
| Q | 165 hCoV-19/England/QEUH-AD23E6/2020\|EPI_ISL_634145\|20 | ···················· | ···················· | ···················· | ···················· | ···················· | ···················· |
| Q | 166 hCoV-19/England/QEUH-AD4830/2020\|EPI_ISL_634146\|20 | ···················· | ···················· | ···················· | ···················· | ···················· | ···················· |
| Q | 167 hCoV-19/England/QEUH-AD2C09/2020\|EPI_ISL_634147\|20 | ···················· | ···················· | ···················· | ···················· | ···················· | ···················· |
| Q | 168 hCoV-19/England/QEUH-AD51CB/2020\|EPI_ISL_634150\|20 | ···················· | ·A·················· | ···················· | ···················· | ···················· | ··A················· |
| Q | 169 hCoV-19/England/QEUH-AD28F6/2020\|EPI_ISL_634151\|20 | ···················· | ···················· | ···················· | ···················· | ······T············· | ···················· |
| Q | 170 hCoV-19/England/QEUH-AD4A1C/2020\|EPI_ISL_634152\|20 | ···················· | ···················· | ···················· | ···················· | ···················· | ···················· |
| Q | 171 hCoV-19/England/QEUH-AD200D/2020\|EPI_ISL_634153\|20 | ···················· | ···················· | ···················· | ···················· | ···················· | ···················· |
| Q | 172 hCoV-19/England/QEUH-AD2B75/2020\|EPI_ISL_634154\|20 | ···················· | ···················· | ···················· | ···················· | ···················· | ···················· |
| Q | 173 hCoV-19/England/QEUH-AD379E/2020\|EPI_ISL_634155\|20 | ···················· | ···················· | ···················· | ···················· | ···················· | ···················· |
| Q | 174 hCoV-19/England/QEUH-AD5116/2020\|EPI_ISL_634157\|20 | ···················· | ···················· | ···················· | ···················· | ···················· | ···················· |
| Q | 175 hCoV-19/England/QEUH-AD5240/2020\|EPI_ISL_634158\|20 | ···················· | ···················· | ···················· | ···················· | ···················· | ···················· |
| Q | 176 hCoV-19/England/QEUH-AD49C4/2020\|EPI_ISL_634159\|20 | ···················· | ···················· | ···················· | ···················· | ···················· | ···················· |
| Q | 177 hCoV-19/England/QEUH-AD1FD4/2020\|EPI_ISL_634160\|20 | ···················· | ···················· | ···················· | ···················· | ···················· | ···················· |
| Q | 178 hCoV-19/England/QEUH-AD202B/2020\|EPI_ISL_634161\|20 | ···················· | ···················· | ···················· | ···················· | ···················· | ···················· |
| Q | 179 hCoV-19/England/QEUH-AD0403/2020\|EPI_ISL_634162\|20 | ···················· | ···················· | ···················· | ···················· | ···················· | ···················· |
| Q | 180 hCoV-19/England/QEUH-AD2D15/2020\|EPI_ISL_634163\|20 | ···················· | ···················· | ···················· | ···················· | ···················· | ···················· |
| Q | 181 hCoV-19/England/QEUH-AD4979/2020\|EPI_ISL_634165\|20 | ···················· | ···················· | ···················· | ···················· | ···················· | ···················· |
| Q | 182 hCoV-19/England/QEUH-AD30EE/2020\|EPI_ISL_634166\|20 | ···················· | ···················· | ···················· | ···················· | ···················· | ···················· |
| Q | 183 hCoV-19/England/QEUH-AD2A0F/2020\|EPI_ISL_634167\|20 | ···················· | ···················· | ···················· | ···················· | ···················· | ···················· |
| Q | 184 hCoV-19/England/QEUH-AD2B48/2020\|EPI_ISL_634168\|20 | ···················· | ···················· | ···················· | ···················· | ···················· | ···················· |
| Q | 185 hCoV-19/England/QEUH-AD20EF/2020\|EPI_ISL_634169\|20 | ···················· | ···················· | ···················· | ···················· | ···················· | ···················· |
| Q | 186 hCoV-19/England/QEUH-AD0C83/2020\|EPI_ISL_634170\|20 | ···················· | ···················· | ···················· | ···················· | ···················· | ···················· |
| Q | 187 hCoV-19/England/QEUH-AD2BEE/2020\|EPI_ISL_634171\|20 | ···················· | ···················· | ···················· | ···················· | ···················· | ···················· |
| Q | 188 hCoV-19/England/QEUH-AD5161/2020\|EPI_ISL_634172\|20 | ···················· | ···················· | ···················· | ···················· | ···················· | ···················· |
| Q | 189 hCoV-19/England/QEUH-AD37CB/2020\|EPI_ISL_634173\|20 | ···················· | ···················· | ···················· | ···················· | ···················· | ···················· |
| Q | 190 hCoV-19/England/QEUH-AD3084/2020\|EPI_ISL_634174\|20 | ···················· | ···················· | ···················· | ···················· | ···················· | ···················· |
| Q | 191 hCoV-19/England/QEUH-AD48D6/2020\|EPI_ISL_634175\|20 | ···················· | ···················· | ···················· | ···················· | ···················· | ···················· |
| Q | 192 hCoV-19/England/QEUH-AD046D/2020\|EPI_ISL_634176\|20 | ···················· | ···················· | ···················· | ···················· | ···················· | ···················· |
| Q | 193 hCoV-19/England/QEUH-AD3D05/2020\|EPI_ISL_634179\|20 | ···················· | ···················· | ···················· | ···················· | ···················· | ···················· |
| Q | 194 hCoV-19/England/QEUH-AD2261/2020\|EPI_ISL_634180\|20 | ···················· | ···················· | ···················· | ···················· | ···················· | ···················· |
| Q | 195 hCoV-19/England/QEUH-AD4645/2020\|EPI_ISL_634181\|20 | ···················· | ···················· | ···················· | ···················· | ···················· | ···················· |
| Q | 196 hCoV-19/England/QEUH-AD341F/2020\|EPI_ISL_634182\|20 | ···················· | ···················· | ···················· | ···················· | ···················· | ···················· |
| Q | 197 hCoV-19/England/QEUH-AD4117/2020\|EPI_ISL_634183\|20 | ···················· | ···················· | ···················· | ···················· | ···················· | ···················· |
| Q | 198 hCoV-19/England/QEUH-AD2A2D/2020\|EPI_ISL_634184\|20 | ···················· | ···················· | ···················· | ···················· | ···················· | ···················· |
| Q | 199 hCoV-19/England/QEUH-AD2498/2020\|EPI_ISL_634185\|20 | ···················· | ···················· | ···················· | ···················· | ···················· | ···················· |
| Q | 200 hCoV-19/England/QEUH-AD49F1/2020\|EPI_ISL_634186\|20 | ···················· | ···················· | ···················· | ···················· | ···················· | ···················· |
| Q | 201 hCoV-19/England/QEUH-AD0CDE/2020\|EPI_ISL_634188\|20 | ···················· | ···················· | ···················· | ···················· | ···················· | ···················· |
| Q | 202 hCoV-19/England/QEUH-AD4627/2020\|EPI_ISL_634189\|20 | ···················· | ···················· | ···················· | ···················· | ···················· | ···················· |
| Q | 203 hCoV-19/England/QEUH-AD484F/2020\|EPI_ISL_634190\|20 | ···················· | ···················· | ···················· | ···················· | ···················· | ···················· |
| Q | 204 hCoV-19/England/QEUH-AD48E5/2020\|EPI_ISL_634191\|20 | ···················· | ···················· | ···················· | ···················· | ···················· | ···················· |
| Q | 205 hCoV-19/England/QEUH-AD51BC/2020\|EPI_ISL_634192\|20 | ···················· | ···················· | ···················· | ···················· | ···················· | ···················· |
| Q | 206 hCoV-19/England/QEUH-AD529B/2020\|EPI_ISL_634193\|20 | ···················· | ···················· | ···················· | ···················· | ···················· | ···················· |
| Q | 207 hCoV-19/England/QEUH-AD3488/2020\|EPI_ISL_634195\|20 | ···················· | ···················· | ···················· | ···················· | ···················· | ···················· |
| Q | 208 hCoV-19/England/QEUH-AD0CB0/2020\|EPI_ISL_634196\|20 | ···················· | ···················· | ···················· | ···················· | ···················· | ···················· |
| Q | 209 hCoV-19/England/QEUH-AD2489/2020\|EPI_ISL_634197\|20 | ···················· | ···················· | ···················· | ···················· | ···················· | ···················· |
| Q | 210 hCoV-19/England/QEUH-AD05A6/2020\|EPI_ISL_634198\|20 | ···················· | ···················· | ···················· | ···················· | ···················· | ···················· |
| Q | 211 hCoV-19/England/QEUH-AD3B92/2020\|EPI_ISL_634199\|20 | ···················· | ···················· | ···················· | ···················· | ···················· | ···················· |
| Q | 212 hCoV-19/USA/CA-ALSR-2382/2020\|EPI_ISL_635451\|2020- | ···················· | ···················· | ···················· | ···················· | ···················· | ···················· |
| Q | 213 hCoV-19/USA/CA-ALSR-2384/2020\|EPI_ISL_635452\|2020- | ···················· | ·A·················· | ···················· | ···················· | ···················· | ···················· |
| Q | 214 hCoV-19/USA/CA-ALSR-2386/2020\|EPI_ISL_635453\|2020- | ···················· | ·A·················· | ···················· | ···················· | ···················· | ···················· |
| Q | 215 hCoV-19/USA/CA-ALSR-2387/2020\|EPI_ISL_635454\|2020- | ···················· | ·A·················· | ···················· | ···················· | ···················· | ···················· |
| Q | 216 hCoV-19/USA/CA-ALSR-2388/2020\|EPI_ISL_635455\|2020- | ···················· | ·A·················· | ···················· | ···················· | ···················· | ···················· |
| Q | 217 hCoV-19/USA/CA-ALSR-2390/2020\|EPI_ISL_635456\|2020- | ···················· | ·A·················· | ···················· | ···················· | ···················· | ···················· |
| Q | 218 hCoV-19/USA/CA-ALSR-2393/2020\|EPI_ISL_635457\|2020- | ···················· | ···················· | ···················· | ···················· | ···················· | ···················· |
| Q | 219 hCoV-19/USA/CA-ALSR-2399/2020\|EPI_ISL_635458\|2020- | ···················· | ···················· | ···················· | ···················· | ···················· | ···················· |
| Q | 220 hCoV-19/USA/CA-ALSR-2400/2020\|EPI_ISL_635459\|2020- | ···················· | ·A·················· | ···················· | ···················· | ···················· | ···················· |
| Q | 221 hCoV-19/USA/CA-ALSR-2404/2020\|EPI_ISL_635460\|2020- | ···················· | ·A·················· | ···················· | ···················· | ···················· | ···················· |
| Q | 222 hCoV-19/USA/CA-ALSR-2406/2020\|EPI_ISL_635462\|2020- | ···················· | ·A·················· | ···················· | ···················· | ···················· | ···················· |
| Q | 223 hCoV-19/USA/CA-ALSR-2407/2020\|EPI_ISL_635463\|2020- | ···················· | ·A·················· | ···················· | ···················· | ···················· | ···················· |
| Q | 224 hCoV-19/USA/CA-ALSR-2410/2020\|EPI_ISL_635466\|2020- | ···················· | ·A·················· | ···················· | ···················· | ···················· | ···················· |
| Q | 225 hCoV-19/USA/CA-ALSR-2412/2020\|EPI_ISL_635467\|2020- | ···················· | ···················· | ···················· | ···················· | ···················· | ···················· |
| Q | 226 hCoV-19/USA/CA-ALSR-2415/2020\|EPI_ISL_635468\|2020- | ···················· | ···················· | ···················· | ···················· | ···················· | ···················· |
| Q | 227 hCoV-19/USA/CA-ALSR-2417/2020\|EPI_ISL_635469\|2020- | ···················· | ·A·················· | ···················· | ···················· | ···················· | ···················· |
| Q | 228 hCoV-19/USA/CA-ALSR-2420/2020\|EPI_ISL_635470\|2020- | ···················· | ·A·················· | ···················· | ···················· | ···················· | ···················· |
| Q | 229 hCoV-19/USA/CA-ALSR-2422/2020\|EPI_ISL_635471\|2020- | ···················· | ·A·················· | ···················· | ···················· | ···················· | ···················· |
| Q | 230 hCoV-19/USA/CA-ALSR-2424/2020\|EPI_ISL_635472\|2020- | ···················· | ·A·················· | ···················· | ···················· | ···················· | ···················· |
| Q | 231 hCoV-19/USA/CA-ALSR-2425/2020\|EPI_ISL_635473\|2020- | ···················· | ···················· | ···················· | ···················· | ···················· | ···················· |

| | | | | | | | |
|---|---|---|---|---|---|---|---|
| Q | 232 hCoV-19/USA/CA-ALSR-2428/2020\|EPI_ISL_635474\|2020- | ···················· | ·A·················· | ····················· | ···················· | ···················· | ···················· |
| Q | 233 hCoV-19/USA/CA-ALSR-2429/2020\|EPI_ISL_635475\|2020- | ···················· | ···················· | ····················· | ···················· | ···················· | ···················· |
| Q | 234 hCoV-19/USA/CA-ALSR-2434/2020\|EPI_ISL_635476\|2020- | ···················· | ·A·················· | ····················· | ···················· | ···················· | ···················· |
| Q | 235 hCoV-19/USA/CA-ALSR-2436/2020\|EPI_ISL_635477\|2020- | ···················· | ·A·················· | ····················· | ···················· | ···················· | ···················· |
| Q | 236 hCoV-19/USA/CA-ALSR-2437/2020\|EPI_ISL_635478\|2020- | ···················· | ···················· | ····················· | ···················· | ···················· | ···················· |
| Q | 237 hCoV-19/Mexico/BCN-ALSR-2446/2020\|EPI_ISL_635480\|2 | ···················· | ·A·················· | ····················· | ···················· | ···················· | ···················· |
| Q | 238 hCoV-19/Mexico/BCN-ALSR-2447/2020\|EPI_ISL_635481\|2 | ···················· | ·A·················· | ····················· | ···················· | ···················· | ···················· |
| Q | 239 hCoV-19/Mexico/BCN-ALSR-2451/2020\|EPI_ISL_635483\|2 | ···················· | ·A·················· | ····················· | ···················· | ···················· | ···················· |
| Q | 240 hCoV-19/Mexico/BCN-ALSR-2452/2020\|EPI_ISL_635484\|2 | ···················· | ···················· | ····················· | ···················· | ···················· | ···················· |
| Q | 241 hCoV-19/Mexico/BCN-ALSR-2462/2020\|EPI_ISL_635486\|2 | ···················· | ·A·················· | ····················· | ···················· | ···················· | ···················· |
| Q | 242 hCoV-19/Mexico/BCN-ALSR-2475/2020\|EPI_ISL_635489\|2 | ···················· | ·A·················· | ····················· | ···················· | ···················· | ···················· |
| Q | 243 hCoV-19/Mexico/BCN-ALSR-2479/2020\|EPI_ISL_635491\|2 | ···················· | ·A·················· | ····················· | ···················· | ···················· | ···················· |
| Q | 244 hCoV-19/Mexico/BCN-ALSR-2480/2020\|EPI_ISL_635492\|2 | ···················· | ·A·················· | ····················· | ···················· | ···················· | ···················· |
| Q | 245 hCoV-19/Mexico/BCN-ALSR-2481/2020\|EPI_ISL_635493\|2 | ···················· | ···················· | ····················· | ···················· | ···················· | ···················· |
| Q | 246 hCoV-19/Mexico/BCN-ALSR-2483/2020\|EPI_ISL_635494\|2 | ···················· | ···················· | ····················· | ···················· | ···················· | ···················· |
| Q | 247 hCoV-19/Mexico/BCN-ALSR-2484/2020\|EPI_ISL_635495\|2 | ···················· | ·A·················· | ····················· | ···················· | ···················· | ···················· |
| Q | 248 hCoV-19/Mexico/BCN-ALSR-2485/2020\|EPI_ISL_635496\|2 | ···················· | ·A·················· | ····················· | ···················· | ···················· | ···················· |
| Q | 249 hCoV-19/Mexico/BCN-ALSR-2487/2020\|EPI_ISL_635497\|2 | ···················· | ·A·················· | ····················· | ···················· | ···················· | ···················· |
| Q | 250 hCoV-19/Mexico/BCN-ALSR-2489/2020\|EPI_ISL_635498\|2 | ···················· | ·A·················· | ····················· | ···················· | ···················· | ···················· |
| Q | 251 hCoV-19/Mexico/BCN-ALSR-2490/2020\|EPI_ISL_635499\|2 | ···················· | ·A·················· | ····················· | ···················· | ···················· | ···················· |
| Q | 252 hCoV-19/India/DL-60932-CSIR-IGIB/2020\|EPI_ISL_6367 | ···················· | ···················· | ····················· | ···················· | ···················· | ···················· |
| Q | 253 hCoV-19/India/UT-50442-CSIR-IGIB/2020\|EPI_ISL_6367 | ···················· | ·A·················· | ····················· | ···················· | ···················· | ···A················ |
| Q | 254 hCoV-19/India/DL-5504375-CSIR-IGIB/2020\|EPI_ISL_63 | ···················· | ·A·················· | ····················· | ···················· | ···················· | ···A················ |
| Q | 255 hCoV-19/India/DL-60935-CSIR-IGIB/2020\|EPI_ISL_6367 | ···················· | ···················· | ····················· | ···················· | ···················· | ···················· |
| Q | 256 hCoV-19/India/DL-60072-CSIR-IGIB/2020\|EPI_ISL_6367 | ···················· | ···················· | ····················· | ···················· | ···················· | ···················· |
| Q | 257 hCoV-19/Wales/PHWC-485A01/2020\|EPI_ISL_628240\|2020 | ···················· | ···················· | ····················· | ···················· | ···················· | ···················· |
| Q | 258 hCoV-19/Wales/PHWC-485A4D/2020\|EPI_ISL_628241\|2020 | ···················· | ···················· | ····················· | ···················· | ···················· | ···················· |
| Q | 259 hCoV-19/Wales/PHWC-485A5C/2020\|EPI_ISL_628242\|2020 | ···················· | ···················· | ····················· | ···················· | ···················· | ···················· |
| Q | 260 hCoV-19/Wales/PHWC-485A6B/2020\|EPI_ISL_628243\|2020 | ···················· | ···················· | ····················· | ···················· | ···················· | ···················· |
| Q | 261 hCoV-19/Wales/PHWC-485A7A/2020\|EPI_ISL_628244\|2020 | ···················· | ···················· | ····················· | ···················· | ···················· | ···················· |
| Q | 262 hCoV-19/Wales/PHWC-485A89/2020\|EPI_ISL_628245\|2020 | ···················· | ···················· | ····················· | ···················· | ···················· | ···················· |
| Q | 263 hCoV-19/Wales/PHWC-485A98/2020\|EPI_ISL_628246\|2020 | ···················· | ···················· | ····················· | ···················· | ···················· | ···················· |
| Q | 264 hCoV-19/Wales/PHWC-485AA7/2020\|EPI_ISL_628247\|2020 | ···················· | ···················· | ····················· | ···················· | ···················· | ···················· |
| Q | 265 hCoV-19/Wales/PHWC-485AB6/2020\|EPI_ISL_628248\|2020 | ···················· | ···················· | ····················· | ···················· | ···················· | ···················· |
| Q | 266 hCoV-19/Wales/PHWC-485AC5/2020\|EPI_ISL_628249\|2020 | ···················· | ···················· | ····················· | ···················· | ···················· | ···················· |
| Q | 267 hCoV-19/Wales/PHWC-485AD4/2020\|EPI_ISL_628250\|2020 | ···················· | ···················· | ····················· | ···················· | ···················· | ···················· |
| Q | 268 hCoV-19/Wales/PHWC-485AE3/2020\|EPI_ISL_628251\|2020 | ···················· | ···················· | ····················· | ···················· | ···················· | ···················· |
| Q | 269 hCoV-19/Wales/PHWC-485B1D/2020\|EPI_ISL_628252\|2020 | ···················· | ···················· | ····················· | ···················· | ···················· | ···················· |
| Q | 270 hCoV-19/Wales/PHWC-485B2C/2020\|EPI_ISL_628253\|2020 | ···················· | ···················· | ····················· | ···················· | ···················· | ···················· |
| Q | 271 hCoV-19/Wales/PHWC-485B3B/2020\|EPI_ISL_628254\|2020 | ···················· | ···················· | ····················· | ···················· | ···················· | ···················· |
| Q | 272 hCoV-19/Wales/PHWC-485B4A/2020\|EPI_ISL_628255\|2020 | ···················· | ···················· | ····················· | ···················· | ···················· | ···················· |
| Q | 273 hCoV-19/Wales/PHWC-485B59/2020\|EPI_ISL_628256\|2020 | ···················· | ···················· | ····················· | ···················· | ···················· | ···················· |
| Q | 274 hCoV-19/Wales/PHWC-485B77/2020\|EPI_ISL_628257\|2020 | ···················· | ···················· | ····················· | ···················· | ···················· | ···················· |
| Q | 275 hCoV-19/Wales/PHWC-485B86/2020\|EPI_ISL_628258\|2020 | ···················· | ···················· | ····················· | ···················· | ···················· | ···················· |
| Q | 277 hCoV-19/Wales/PHWC-485BA4/2020\|EPI_ISL_628260\|2020 | ···················· | ···················· | ····················· | ···················· | ···················· | ···················· |
| Q | 278 hCoV-19/Wales/PHWC-485BC2/2020\|EPI_ISL_628261\|2020 | ···················· | ···················· | ····················· | ···················· | ···················· | ···················· |
| Q | 279 hCoV-19/Wales/PHWC-485C65/2020\|EPI_ISL_628270\|2020 | ···················· | ···················· | ····················· | ···················· | ···················· | ···················· |
| Q | 280 hCoV-19/Wales/PHWC-485CCF/2020\|EPI_ISL_628271\|2020 | ···················· | ···················· | ····················· | ···················· | ···················· | ···················· |
| Q | 282 hCoV-19/Wales/PHWC-485D08/2020\|EPI_ISL_628273\|2020 | ···················· | ···················· | ····················· | ···················· | ···················· | ···················· |
| Q | 283 hCoV-19/Wales/PHWC-485D26/2020\|EPI_ISL_628274\|2020 | ···················· | ···················· | ····················· | ···················· | ···················· | ···················· |
| Q | 284 hCoV-19/Wales/PHWC-485D35/2020\|EPI_ISL_628275\|2020 | ···················· | ···················· | ····················· | ···················· | ···················· | ···················· |
| Q | 285 hCoV-19/Wales/PHWC-485D44/2020\|EPI_ISL_628276\|2020 | ···················· | ···················· | ····················· | ···················· | ···················· | ···················· |
| Q | 286 hCoV-19/Wales/PHWC-485D62/2020\|EPI_ISL_628277\|2020 | ···················· | ···················· | ····················· | ···················· | ···················· | ···················· |
| Q | 287 hCoV-19/Wales/PHWC-485D80/2020\|EPI_ISL_628278\|2020 | ···················· | ···················· | ····················· | ···················· | ···················· | ···················· |
| Q | 288 hCoV-19/Wales/PHWC-485DAE/2020\|EPI_ISL_628279\|2020 | ···················· | ···················· | ····················· | ···················· | ···················· | ···················· |
| Q | 290 hCoV-19/Wales/PHWC-485DCC/2020\|EPI_ISL_628281\|2020 | ···················· | ···················· | ····················· | ···················· | ···················· | ···················· |
| Q | 291 hCoV-19/Wales/PHWC-485DDB/2020\|EPI_ISL_628282\|2020 | ···················· | ···················· | ····················· | ···················· | ···················· | ···················· |
| Q | 292 hCoV-19/Wales/PHWC-485E05/2020\|EPI_ISL_628283\|2020 | ···················· | ···················· | ····················· | ···················· | ···················· | ···················· |
| Q | 293 hCoV-19/Wales/PHWC-485E23/2020\|EPI_ISL_628284\|2020 | ···················· | ···················· | ····················· | ···················· | ···················· | ···················· |
| Q | 294 hCoV-19/Wales/PHWC-485E41/2020\|EPI_ISL_628285\|2020 | ···················· | ···················· | ····················· | ···················· | ···················· | ···················· |
| Q | 295 hCoV-19/Wales/PHWC-485E50/2020\|EPI_ISL_628286\|2020 | ···················· | ···················· | ····················· | ···················· | ···················· | ···················· |
| Q | 296 hCoV-19/Wales/PHWC-485E6F/2020\|EPI_ISL_628287\|2020 | ···················· | ···················· | ····················· | ···················· | ···················· | ···················· |
| Q | 297 hCoV-19/Wales/PHWC-485E8D/2020\|EPI_ISL_628288\|2020 | ···················· | ···················· | ····················· | ···················· | ···················· | ···················· |
| Q | 298 hCoV-19/Wales/PHWC-485E9C/2020\|EPI_ISL_628289\|2020 | ···················· | ···················· | ····················· | ···················· | ···················· | ···················· |
| Q | 299 hCoV-19/Wales/PHWC-485EBA/2020\|EPI_ISL_628290\|2020 | ···················· | ···················· | ····················· | ···················· | ···················· | ···················· |
| Q | 300 hCoV-19/Wales/PHWC-485EC9/2020\|EPI_ISL_628291\|2020 | ···················· | ···················· | ····················· | ···················· | ···················· | ···················· |
| Q | 301 hCoV-19/Wales/PHWC-485ED8/2020\|EPI_ISL_628292\|2020 | ···················· | ···················· | ····················· | ···················· | ···················· | ···················· |
| Q | 302 hCoV-19/Wales/PHWC-485F20/2020\|EPI_ISL_628293\|2020 | ···················· | ···················· | ····················· | ···················· | ···················· | ···················· |
| Q | 303 hCoV-19/Wales/PHWC-485F5D/2020\|EPI_ISL_628294\|2020 | ···················· | ···················· | ····················· | ···················· | ···················· | ···················· |
| Q | 304 hCoV-19/Wales/PHWC-485F6C/2020\|EPI_ISL_628295\|2020 | ···················· | ···················· | ····················· | ···················· | ···················· | ···················· |
| Q | 305 hCoV-19/Wales/PHWC-485F7B/2020\|EPI_ISL_628296\|2020 | ···················· | ···················· | ····················· | ···················· | ···················· | ···················· |
| Q | 306 hCoV-19/Wales/PHWC-485F8A/2020\|EPI_ISL_628297\|2020 | ···················· | ···················· | ····················· | ···················· | ···················· | ···················· |
| Q | 307 hCoV-19/Wales/PHWC-485F99/2020\|EPI_ISL_628298\|2020 | ···················· | ···················· | ····················· | ···················· | ···················· | ···················· |
| Q | 308 hCoV-19/Wales/PHWC-485FB7/2020\|EPI_ISL_628299\|2020 | ···················· | ···················· | ····················· | ···················· | ···················· | ···················· |
| Q | 309 hCoV-19/Wales/PHWC-485FD5/2020\|EPI_ISL_628300\|2020 | ···················· | ···················· | ····················· | ···················· | ···················· | ···················· |
| Q | 310 hCoV-19/Wales/PHWC-48600E/2020\|EPI_ISL_628301\|2020 | ···················· | ···················· | ····················· | ···················· | ···················· | ···················· |
| Q | 311 hCoV-19/Wales/PHWC-48603B/2020\|EPI_ISL_628302\|2020 | ···················· | ···················· | ····················· | ···················· | ···················· | ···················· |
| Q | 312 hCoV-19/Wales/PHWC-48604A/2020\|EPI_ISL_628303\|2020 | ···················· | ···················· | ····················· | ···················· | ···················· | ···················· |
| Q | 313 hCoV-19/Wales/PHWC-486059/2020\|EPI_ISL_628304\|2020 | ···················· | ···················· | ····················· | ···················· | ···················· | ···················· |
| Q | 314 hCoV-19/Wales/PHWC-486077/2020\|EPI_ISL_628305\|2020 | ···················· | ···················· | ····················· | ···················· | ···················· | ···················· |

| | | | | | | | |
|---|---|---|---|---|---|---|---|
| Q | 315 hCoV-19/Wales/PHWC-486095/2020\|EPI_ISL_628306\|2020 | ···················· | ···················· | ····················· | ···················· | ···················· | ···················· |
| Q | 317 hCoV-19/Wales/PHWC-4860E0/2020\|EPI_ISL_628308\|2020 | ···················· | ···················· | ····················· | ···················· | ···················· | ···················· |
| Q | 318 hCoV-19/Wales/PHWC-4860FF/2020\|EPI_ISL_628309\|2020 | ···················· | ···················· | ····················· | ···················· | ···················· | ···················· |
| Q | 319 hCoV-19/Wales/PHWC-486129/2020\|EPI_ISL_628310\|2020 | ···················· | ···················· | ····················· | ···················· | ···················· | ···················· |
| Q | 320 hCoV-19/Wales/PHWC-486147/2020\|EPI_ISL_628311\|2020 | ···················· | ···················· | ····················· | ···················· | ···················· | ···················· |
| Q | 321 hCoV-19/England/PORT-2D31A7/2020\|EPI_ISL_628312\|20 | ···················· | ···················· | ····················· | ···················· | ···················· | ···················· |
| Q | 322 hCoV-19/England/PORT-2D31B6/2020\|EPI_ISL_628313\|20 | ···················· | ···················· | ····················· | ···················· | ···················· | ···················· |
| Q | 323 hCoV-19/England/PORT-2D31C5/2020\|EPI_ISL_628314\|20 | ···················· | ···················· | ····················· | ···················· | ···················· | ···················· |
| Q | 324 hCoV-19/England/PORT-2D31E3/2020\|EPI_ISL_628315\|20 | ···················· | ···················· | ····················· | ···················· | ···················· | ···················· |
| Q | 325 hCoV-19/England/PORT-2D320E/2020\|EPI_ISL_628316\|20 | ···················· | ···················· | ····················· | ···················· | ···················· | ···················· |
| Q | 326 hCoV-19/England/PORT-2D322C/2020\|EPI_ISL_628317\|20 | ···················· | ···················· | ····················· | ···················· | ···················· | ···················· |
| Q | 327 hCoV-19/England/PORT-2D3259/2020\|EPI_ISL_628318\|20 | ···················· | ···················· | ····················· | ···················· | ···················· | ···················· |
| Q | 328 hCoV-19/England/PORT-2D3277/2020\|EPI_ISL_628319\|20 | ···················· | ···················· | ····················· | ···················· | ···················· | ···················· |
| Q | 329 hCoV-19/England/QEUH-A322AB/2020\|EPI_ISL_628340\|20 | ···················· | ···················· | ····················· | ···················· | ···················· | ···················· |
| Q | 330 hCoV-19/Scotland/QEUH-A322BA/2020\|EPI_ISL_628341\|2 | ···················· | ···················· | ····················· | ···················· | ···················· | ···················· |
| Q | 331 hCoV-19/England/QEUH-A322C9/2020\|EPI_ISL_628342\|20 | ···················· | ···················· | ····················· | ···················· | ···················· | ···················· |
| Q | 332 hCoV-19/Scotland/QEUH-A322D8/2020\|EPI_ISL_628343\|2 | ···················· | ···················· | ····················· | ···················· | ···················· | ···················· |
| Q | 333 hCoV-19/England/QEUH-A322E7/2020\|EPI_ISL_628344\|20 | ···················· | ···················· | ····················· | ···················· | ···················· | ···················· |
| Q | 334 hCoV-19/England/QEUH-A32302/2020\|EPI_ISL_628345\|20 | ···················· | ···················· | ····················· | ···················· | ···················· | ···················· |
| Q | 335 hCoV-19/Scotland/QEUH-A32320/2020\|EPI_ISL_628346\|2 | ···················· | ···················· | ····················· | ···················· | ···················· | ···················· |
| Q | 336 hCoV-19/England/MILK-AC924F/2020\|EPI_ISL_629571\|20 | ···················· | ···················· | ····················· | ···················· | ···················· | ···················· |
| Q | 337 hCoV-19/England/MILK-AC8E00/2020\|EPI_ISL_629574\|20 | ···················· | ·A·················· | ····················· | ···················· | ···················· | ···A················ |
| Q | 338 hCoV-19/England/MILK-ACEF16/2020\|EPI_ISL_629575\|20 | ···················· | ···················· | ····················· | ···················· | ····T··············· | ···················· |
| Q | 339 hCoV-19/England/MILK-ACE268/2020\|EPI_ISL_629576\|20 | ···················· | ···················· | ····················· | ···················· | ···················· | ···················· |
| Q | 340 hCoV-19/England/MILK-AC81BC/2020\|EPI_ISL_629577\|20 | ···················· | ···················· | ····················· | ···················· | ···················· | ···················· |
| Q | 341 hCoV-19/England/MILK-AC7EE3/2020\|EPI_ISL_629578\|20 | ···················· | ···················· | ····················· | ···················· | ···················· | ···················· |
| Q | 342 hCoV-19/England/MILK-AC82B9/2020\|EPI_ISL_629579\|20 | ···················· | ···················· | ····················· | ···················· | ···················· | ···················· |
| Q | 343 hCoV-19/England/MILK-AC828C/2020\|EPI_ISL_629582\|20 | ···················· | ···················· | ····················· | ···················· | ···················· | ···················· |
| Q | 344 hCoV-19/England/MILK-ACF1B5/2020\|EPI_ISL_629584\|20 | ···················· | ···················· | ····················· | ···················· | ···················· | ···················· |
| Q | 345 hCoV-19/England/MILK-ACF7E0/2020\|EPI_ISL_629585\|20 | ···················· | ···················· | ····················· | ···················· | ···················· | ···················· |
| Q | 346 hCoV-19/England/MILK-AC8F0D/2020\|EPI_ISL_629586\|20 | ···················· | ···················· | ····················· | ···················· | ···················· | ···················· |
| Q | 347 hCoV-19/England/MILK-ACEBA9/2020\|EPI_ISL_629587\|20 | ···················· | ···················· | ····················· | ···················· | ···················· | ···················· |
| Q | 348 hCoV-19/England/MILK-AC8091/2020\|EPI_ISL_629588\|20 | ···················· | ···················· | ····················· | ···················· | ···················· | ···················· |
| Q | 349 hCoV-19/England/MILK-ACFA14/2020\|EPI_ISL_629589\|20 | ···················· | ···················· | ····················· | ···················· | ···················· | ···················· |
| Q | 350 hCoV-19/England/MILK-AC7F68/2020\|EPI_ISL_629609\|20 | ···················· | ···················· | ····················· | ···················· | ······T············· | ···················· |
| Q | 351 hCoV-19/England/MILK-AC8F67/2020\|EPI_ISL_629610\|20 | ···················· | ···················· | ····················· | ···················· | ···················· | ···················· |
| Q | 352 hCoV-19/England/MILK-ACF14C/2020\|EPI_ISL_629611\|20 | ···················· | ···················· | ····················· | ···················· | ···················· | ···················· |
| Q | 353 hCoV-19/England/MILK-ACF1C4/2020\|EPI_ISL_629612\|20 | ···················· | ···················· | ····················· | ···················· | ···················· | ···················· |
| Q | 354 hCoV-19/England/MILK-AC9212/2020\|EPI_ISL_629613\|20 | ···················· | ···················· | ····················· | ···················· | ···················· | ···················· |
| Q | 355 hCoV-19/England/MILK-ACE58D/2020\|EPI_ISL_629614\|20 | ···················· | ···················· | ····················· | ···················· | ······T············· | ···················· |
| Q | 356 hCoV-19/England/MILK-ACF0F4/2020\|EPI_ISL_629615\|20 | ···················· | ···················· | ····················· | ···················· | ···················· | ···················· |
| Q | 357 hCoV-19/England/MILK-AC8A1B/2020\|EPI_ISL_629616\|20 | ···················· | ···················· | ····················· | ···················· | ···················· | ···················· |
| Q | 358 hCoV-19/England/MILK-ACE70F/2020\|EPI_ISL_629617\|20 | ···················· | ···················· | ····················· | ···················· | ···················· | ···················· |
| Q | 359 hCoV-19/England/MILK-AC8046/2020\|EPI_ISL_629618\|20 | ···················· | ···················· | ····················· | ···················· | ···················· | ···················· |
| Q | 360 hCoV-19/England/MILK-AC7F77/2020\|EPI_ISL_629619\|20 | ···················· | ···················· | ····················· | ···················· | ···················· | ···················· |
| Q | 361 hCoV-19/England/MILK-AC8556/2020\|EPI_ISL_629621\|20 | ···················· | ···················· | ····················· | ···················· | ···················· | ···················· |
| Q | 362 hCoV-19/England/MILK-AC81CB/2020\|EPI_ISL_629622\|20 | ···················· | ···················· | ····················· | ···················· | ···················· | ···················· |
| Q | 363 hCoV-19/England/MILK-AC90DC/2020\|EPI_ISL_629623\|20 | ···················· | ···················· | ····················· | ···················· | ···················· | ···················· |
| Q | 364 hCoV-19/England/MILK-AC891E/2020\|EPI_ISL_629624\|20 | ···················· | ···················· | ····················· | ···················· | ···················· | ···················· |
| Q | 365 hCoV-19/England/MILK-ACE67B/2020\|EPI_ISL_629625\|20 | ···················· | ···················· | ····················· | ···················· | ···················· | ···················· |
| Q | 366 hCoV-19/England/MILK-AC800A/2020\|EPI_ISL_629626\|20 | ···················· | ···················· | ····················· | ···················· | ···················· | ···················· |
| Q | 367 hCoV-19/England/MILK-AC8073/2020\|EPI_ISL_629627\|20 | ···················· | ···················· | ····················· | ···················· | ···················· | ···················· |
| Q | 368 hCoV-19/England/MILK-ACEF70/2020\|EPI_ISL_629628\|20 | ···················· | ···················· | ····················· | ···················· | ···················· | ···················· |
| Q | 369 hCoV-19/England/MILK-ACF12E/2020\|EPI_ISL_629629\|20 | ···················· | ···················· | ····················· | ···················· | ···················· | ···················· |
| Q | 370 hCoV-19/England/MILK-ACEE73/2020\|EPI_ISL_629630\|20 | ···················· | ···················· | ····················· | ···················· | ···················· | ···················· |
| Q | 371 hCoV-19/England/MILK-ACE66C/2020\|EPI_ISL_629631\|20 | ···················· | ····G··············· | ····················· | ···················· | ···················· | ···················· |
| Q | 372 hCoV-19/England/MILK-ACEEEC/2020\|EPI_ISL_629634\|20 | ···················· | ···················· | ····················· | ···················· | ···················· | ···················· |
| Q | 373 hCoV-19/England/MILK-ACF443/2020\|EPI_ISL_629635\|20 | ···················· | ···················· | ····················· | ···················· | ···················· | ···················· |
| Q | 374 hCoV-19/England/MILK-AC890F/2020\|EPI_ISL_629636\|20 | ···················· | ···················· | ····················· | ···················· | ···················· | ···················· |
| Q | 375 hCoV-19/England/MILK-AC84C2/2020\|EPI_ISL_629637\|20 | ···················· | ···················· | ····················· | ···················· | ···················· | ···················· |
| Q | 376 hCoV-19/England/MILK-AC8055/2020\|EPI_ISL_629639\|20 | ···················· | ···················· | ····················· | ···················· | ···················· | ···················· |
| Q | 377 hCoV-19/England/MILK-ACE2E0/2020\|EPI_ISL_629640\|20 | ···················· | ···················· | ····················· | ···················· | ······T············· | ···················· |
| Q | 378 hCoV-19/England/MILK-ACEFAD/2020\|EPI_ISL_629641\|20 | ···················· | ···················· | ····················· | ···················· | ···················· | ···················· |
| Q | 379 hCoV-19/England/MILK-ACF249/2020\|EPI_ISL_629642\|20 | ···················· | ···················· | ····················· | ···················· | ···················· | ···················· |
| Q | 380 hCoV-19/England/MILK-ACE699/2020\|EPI_ISL_629644\|20 | ···················· | ···················· | ····················· | ···················· | ···················· | ···················· |
| Q | 381 hCoV-19/England/MILK-AC931F/2020\|EPI_ISL_629646\|20 | ···················· | ···················· | ····················· | ···················· | ···················· | ···················· |
| Q | 382 hCoV-19/England/MILK-ACF16A/2020\|EPI_ISL_629647\|20 | ···················· | ···················· | ····················· | ···················· | ···················· | ···················· |
| Q | 383 hCoV-19/England/MILK-ACEB6D/2020\|EPI_ISL_629648\|20 | ···················· | ···················· | ····················· | ···················· | ···················· | ···················· |
| Q | 384 hCoV-19/England/MILK-AC8BAF/2020\|EPI_ISL_629649\|20 | ···················· | ···················· | ····················· | ···················· | ······T············· | ···················· |
| Q | 385 hCoV-19/England/MILK-AC9124/2020\|EPI_ISL_629670\|20 | ···················· | ···················· | ····················· | ···················· | ···················· | ···················· |
| Q | 386 hCoV-19/England/MILK-ACEE28/2020\|EPI_ISL_629671\|20 | ···················· | ···················· | ····················· | ···················· | ···················· | ···················· |
| Q | 387 hCoV-19/England/MILK-ACE9BE/2020\|EPI_ISL_629673\|20 | ···················· | ···················· | ····················· | ···················· | ···················· | ···················· |
| Q | 388 hCoV-19/England/MILK-AC8231/2020\|EPI_ISL_629674\|20 | ···················· | ···················· | ····················· | ···················· | ···················· | ···················· |
| Q | 389 hCoV-19/England/MILK-ACE63F/2020\|EPI_ISL_629675\|20 | ···················· | ···················· | ····················· | ···················· | ···················· | ···················· |
| Q | 390 hCoV-19/England/MILK-ACF2D0/2020\|EPI_ISL_629676\|20 | ···················· | ···················· | ····················· | ···················· | ···················· | ···················· |
| Q | 391 hCoV-19/England/MILK-AC8529/2020\|EPI_ISL_629677\|20 | ···················· | ···················· | ····················· | ···················· | ···················· | ···················· |
| Q | 392 hCoV-19/England/ALDP-A678F6/2020\|EPI_ISL_630555\|20 | ···················· | ···················· | ····················· | ···················· | ···················· | ···················· |
| Q | 393 hCoV-19/England/ALDP-A68664/2020\|EPI_ISL_630556\|20 | ···················· | ···················· | ····················· | ···················· | ···················· | ···················· |
| Q | 394 hCoV-19/England/ALDP-A67A96/2020\|EPI_ISL_630557\|20 | ···················· | ···················· | ····················· | ···················· | ···················· | ···················· |
| Q | 395 hCoV-19/England/ALDP-A672AD/2020\|EPI_ISL_630558\|20 | ···················· | ···················· | ····················· | ···················· | ···················· | ···················· |

```
Q 396 hCoV-19/England/ALDP-A66C64/2020|EPI_ISL_630559|20 .................... .................... .................... .................... .................... ....................
Q 397 hCoV-19/England/ALDP-A67C27/2020|EPI_ISL_630560|20 .................... .................... .................... .................... .................... ....................
Q 398 hCoV-19/England/ALDP-A668AC/2020|EPI_ISL_630561|20 .................... .................... .................... .................... .................... ....................
Q 399 hCoV-19/England/ALDP-A67911/2020|EPI_ISL_630562|20 .................... .A.................. .................... .................... .................... ....................
Q 400 hCoV-19/England/ALDP-A668E8/2020|EPI_ISL_630563|20 .................... .................... .................... .................... .................... ....................
Q 401 hCoV-19/England/ALDP-A66EAA/2020|EPI_ISL_630564|20 .................... .................... .................... .................... .................... ....................
Q 402 hCoV-19/England/ALDP-A67629/2020|EPI_ISL_630566|20 .................... .................... .................... .................... .................... ....................
Q 403 hCoV-19/England/ALDP-A67119/2020|EPI_ISL_630567|20 .................... .................... .................... .................... .................... ....................
Q 404 hCoV-19/England/ALDP-A66B67/2020|EPI_ISL_630568|20 .................... .................... .................... .................... .................... ....................
Q 405 hCoV-19/England/ALDP-A67243/2020|EPI_ISL_630569|20 .................... .................... .................... .................... .................... ....................
Q 406 hCoV-19/England/ALDP-A673C8/2020|EPI_ISL_630570|20 .................... .................... .................... .................... .................... ....................
Q 407 hCoV-19/England/ALDP-A66F98/2020|EPI_ISL_630571|20 .................... .................... .................... .................... .................... ....................
Q 408 hCoV-19/England/ALDP-A6793F/2020|EPI_ISL_630572|20 .................... .................... .................... .................... .................... ....................
Q 409 hCoV-19/England/ALDP-A673B9/2020|EPI_ISL_630573|20 .................... .................... .................... .................... .................... ....................
Q 410 hCoV-19/England/ALDP-A674E3/2020|EPI_ISL_630574|20 .................... .................... .................... .................... .................... ....................
Q 411 hCoV-19/England/ALDP-A67656/2020|EPI_ISL_630575|20 .................... .................... .................... .................... .................... ....................
Q 412 hCoV-19/England/ALDP-A66CCE/2020|EPI_ISL_630584|20 .................... .................... .................... .................... .................... ....................
Q 413 hCoV-19/England/ALDP-A66806/2020|EPI_ISL_630585|20 .................... .................... .................... .................... .................... ....................
Q 414 hCoV-19/England/ALDP-A67683/2020|EPI_ISL_630588|20 .................... .................... .................... .................... .................... ....................
Q 415 hCoV-19/England/ALDP-A6702B/2020|EPI_ISL_630589|20 .................... .................... .................... .................... .................... ....................
Q 416 hCoV-19/England/ALDP-A6860A/2020|EPI_ISL_630590|20 .................... .................... .................... .................... .................... ....................
Q 417 hCoV-19/England/ALDP-A67647/2020|EPI_ISL_630591|20 .................... .................... .................... .................... .................... ....................
Q 418 hCoV-19/England/ALDP-A6738C/2020|EPI_ISL_630592|20 .................... .................... .................... .................... .................... ....................
Q 419 hCoV-19/England/ALDP-A67304/2020|EPI_ISL_630593|20 .................... .................... .................... .................... .................... ....................
Q 420 hCoV-19/England/ALDP-A679E4/2020|EPI_ISL_630594|20 .................... .................... .................... .................... .................... ....................
Q 421 hCoV-19/England/ALDP-A67049/2020|EPI_ISL_630595|20 .................... .................... .................... .................... .................... ....................
Q 422 hCoV-19/England/ALDP-A677F9/2020|EPI_ISL_630596|20 .................... .................... .................... .................... .................... ....................
Q 423 hCoV-19/England/ALDP-A66EB9/2020|EPI_ISL_630597|20 .................... .................... .................... .................... .................... ....................
Q 424 hCoV-19/England/ALDP-A68716/2020|EPI_ISL_630598|20 .................... .................... .................... .................... .................... ....................
Q 425 hCoV-19/England/ALDP-A67586/2020|EPI_ISL_630599|20 .................... .................... .................... .................... .................... ....................
Q 426 hCoV-19/England/ALDP-A6687F/2020|EPI_ISL_630601|20 .................... .................... .................... .................... .................... ....................
Q 427 hCoV-19/England/ALDP-A678C9/2020|EPI_ISL_630602|20 .................... .................... .................... .................... .................... ....................
Q 428 hCoV-19/England/ALDP-A678AB/2020|EPI_ISL_630603|20 .................... .................... .................... .................... .................... ....................
Q 429 hCoV-19/England/ALDP-A67902/2020|EPI_ISL_630604|20 .................... .................... .................... .................... .................... ....................
Q 430 hCoV-19/England/ALDP-A66D52/2020|EPI_ISL_630605|20 .................... .................... .................... .................... .................... ....................
Q 431 hCoV-19/England/ALDP-A675B3/2020|EPI_ISL_630606|20 .................... .................... .................... .................... .................... ....................
Q 432 hCoV-19/England/ALDP-A668CA/2020|EPI_ISL_630607|20 .................... .A.................. .................... .................... .................... ..A.................
Q 433 hCoV-19/England/ALDP-A66F01/2020|EPI_ISL_630608|20 .................... .................... .................... .................... .................... ....................
Q 434 hCoV-19/England/ALDP-A67C54/2020|EPI_ISL_630610|20 .................... .................... .................... .................... .................... ....................
Q 435 hCoV-19/England/ALDP-A66921/2020|EPI_ISL_630611|20 .................... .................... .................... .................... .................... ....................
Q 436 hCoV-19/England/ALDP-A6728F/2020|EPI_ISL_630612|20 .................... .................... .................... .................... .................... ....................
Q 437 hCoV-19/England/ALDP-A689B6/2020|EPI_ISL_630613|20 .................... .................... .................... .................... .................... ....................
Q 438 hCoV-19/England/ALDP-A67805/2020|EPI_ISL_630614|20 .................... .................... .................... .................... .................... ....................
Q 439 hCoV-19/England/ALDP-A67AF0/2020|EPI_ISL_630615|20 .................... .................... .................... .................... .................... ....................
Q 440 hCoV-19/England/ALDP-A67717/2020|EPI_ISL_630616|20 .................... .................... .................... .................... .................... ....................
Q 441 hCoV-19/England/ALDP-A686BF/2020|EPI_ISL_630617|20 .................... .................... .................... .................... .................... ....................
Q 442 hCoV-19/England/ALDP-A67182/2020|EPI_ISL_630618|20 .................... .A.................. .................... .................... .................... ....................
Q 443 hCoV-19/England/ALDP-A6739B/2020|EPI_ISL_630619|20 .................... .................... .................... .................... .................... ....................
Q 444 hCoV-19/England/ALDP-A667CD/2020|EPI_ISL_630620|20 .................... .................... .................... .................... .................... ....................
Q 445 hCoV-19/England/ALDP-A6751D/2020|EPI_ISL_630622|20 .................... .................... .................... .................... .................... ....................
Q 446 hCoV-19/England/ALDP-A66EC8/2020|EPI_ISL_630623|20 .................... .................... .................... .................... .................... ....................
Q 447 hCoV-19/England/ALDP-A66C46/2020|EPI_ISL_630624|20 .................... .................... .................... .................... .................... ....................
Q 448 hCoV-19/England/ALDP-A66A6A/2020|EPI_ISL_630625|20 .................... .................... .................... .................... .................... ....................
Q 449 hCoV-19/England/ALDP-A66AB5/2020|EPI_ISL_630626|20 .................... .................... .................... .................... .................... ....................
Q 450 hCoV-19/England/ALDP-A67B0C/2020|EPI_ISL_630627|20 .................... .................... .................... .................... .................... ....................
Q 451 hCoV-19/England/ALDP-A66CFB/2020|EPI_ISL_630628|20 .................... .................... .................... .................... .................... ....................
Q 452 hCoV-19/England/ALDP-A66903/2020|EPI_ISL_630629|20 .................... .................... .................... .................... .................... ....................
Q 453 hCoV-19/England/ALDP-A668F7/2020|EPI_ISL_630630|20 .................... .A.................. .................... .................... .................... ....................
Q 454 hCoV-19/England/ALDP-A674C5/2020|EPI_ISL_630631|20 .................... .................... .................... .................... .................... ....................
Q 455 hCoV-19/England/ALDP-A68DAB/2020|EPI_ISL_630654|20 .................... .................... .................... .................... .................... ....................
Q 456 hCoV-19/England/ALDP-A67410/2020|EPI_ISL_630655|20 .................... .................... .................... .................... .................... ....................
Q 457 hCoV-19/England/ALDP-A66BD0/2020|EPI_ISL_630656|20 .................... .................... .................... .................... .................... ....................
Q 458 hCoV-19/USA/NY-NYCPHL-000403/2020|EPI_ISL_631887|2 .................... .................... .................... .................... .................... ....................
Q 459 hCoV-19/USA/NY-NYCPHL-000404/2020|EPI_ISL_631888|2 .................... .................... .................... .................... .................... ....................
Q 460 hCoV-19/USA/NY-NYCPHL-000405/2020|EPI_ISL_631889|2 .................... .................... .................... .................... .................... ....................
Q 461 hCoV-19/USA/NY-NYCPHL-000406/2020|EPI_ISL_631890|2 .................... .................... .................... .................... .................... ....................
Q 462 hCoV-19/USA/NY-NYCPHL-000407/2020|EPI_ISL_631891|2 .................... .................... .................... .................... .................... ....................
Q 463 hCoV-19/USA/NY-NYCPHL-000408/2020|EPI_ISL_631892|2 .................... .................... .................... .................... .................... ....................
Q 464 hCoV-19/USA/NY-NYCPHL-000409/2020|EPI_ISL_631893|2 .................... .................... .................... .................... .................... ....................
Q 465 hCoV-19/USA/NY-NYCPHL-000410/2020|EPI_ISL_631894|2 .................... .................... .................... .................... .................... ....................
Q 466 hCoV-19/USA/NY-NYCPHL-000411/2020|EPI_ISL_631895|2 .................... .................... .................... .................... .................... ....................
Q 467 hCoV-19/USA/NY-NYCPHL-000412/2020|EPI_ISL_631896|2 .................... .................... .................... .................... .................... ....................
Q 468 hCoV-19/USA/NY-NYCPHL-000413/2020|EPI_ISL_631897|2 .................... .................... .................... .................... .................... ....................
Q 469 hCoV-19/USA/NY-NYCPHL-000414/2020|EPI_ISL_631898|2 .................... .................... .................... .................... .................... ....................
Q 470 hCoV-19/USA/NY-NYCPHL-000446/2020|EPI_ISL_631920|2 .................... .................... .................... .................... .................... ....................
Q 471 hCoV-19/USA/NY-NYCPHL-000447/2020|EPI_ISL_631921|2 .................... .................... .................... .................... .................... ....................
Q 472 hCoV-19/USA/NY-NYCPHL-000448/2020|EPI_ISL_631922|2 .................... .................... .................... .................... .................... ....................
Q 473 hCoV-19/USA/NY-NYCPHL-000449/2020|EPI_ISL_631923|2 .................... .................... .................... .................... .................... ....................
Q 474 hCoV-19/USA/NY-NYCPHL-000450/2020|EPI_ISL_631924|2 .................... .................... .................... .................... .................... ....................
Q 475 hCoV-19/USA/NY-NYCPHL-000451/2020|EPI_ISL_631925|2 .................... .................... .................... .................... .................... ....................
```

| | | | | | | | |
|---|---|---|---|---|---|---|---|
| Q | 476 hCoV-19/USA/NY-NYCPHL-000452/2020\|EPI_ISL_631926\|2 | ···················· | ···················· | ····················· | ···················· | ···················· | ···················· |
| Q | 477 hCoV-19/USA/NY-NYCPHL-000453/2020\|EPI_ISL_631927\|2 | ···················· | ···················· | ····················· | ···················· | ···················· | ···················· |
| Q | 478 hCoV-19/USA/NY-NYCPHL-000454/2020\|EPI_ISL_631928\|2 | ···················· | ···················· | ····················· | ···················· | ···················· | ···················· |
| Q | 479 hCoV-19/USA/NY-NYCPHL-000456/2020\|EPI_ISL_631929\|2 | ···················· | ···················· | ····················· | ···················· | ···················· | ···················· |
| Q | 480 hCoV-19/USA/NY-NYCPHL-000458/2020\|EPI_ISL_631931\|2 | ···················· | ···················· | ····················· | ···················· | ···················· | ···················· |
| Q | 481 hCoV-19/USA/NY-NYCPHL-000461/2020\|EPI_ISL_631932\|2 | ···················· | ···················· | ····················· | ···················· | ···················· | ···················· |
| Q | 482 hCoV-19/USA/NY-NYCPHL-000463/2020\|EPI_ISL_631933\|2 | ···················· | ···················· | ····················· | ···················· | ···················· | ···················· |
| Q | 483 hCoV-19/USA/NY-NYCPHL-000464/2020\|EPI_ISL_631934\|2 | ···················· | ···················· | ····················· | ···················· | ···················· | ···················· |
| Q | 484 hCoV-19/USA/NY-NYCPHL-000465/2020\|EPI_ISL_631935\|2 | ···················· | ···················· | ····················· | ···················· | ···················· | ···················· |
| Q | 485 hCoV-19/USA/NY-NYCPHL-000466/2020\|EPI_ISL_631936\|2 | ···················· | ···················· | ····················· | ···················· | ···················· | ···················· |
| Q | 486 hCoV-19/USA/NY-NYCPHL-000467/2020\|EPI_ISL_631937\|2 | ···················· | ···················· | ····················· | ···················· | ···················· | ···················· |
| Q | 487 hCoV-19/USA/NY-NYCPHL-000468/2020\|EPI_ISL_631938\|2 | ···················· | ···················· | ····················· | ···················· | ···················· | ···················· |
| Q | 488 hCoV-19/USA/NY-NYCPHL-000469/2020\|EPI_ISL_631939\|2 | ···················· | ···················· | ····················· | ···················· | ···················· | ···················· |
| Q | 489 hCoV-19/USA/NY-NYCPHL-000470/2020\|EPI_ISL_631940\|2 | ···················· | ···················· | ····················· | ···················· | ···················· | ···················· |
| Q | 490 hCoV-19/USA/NY-NYCPHL-000473/2020\|EPI_ISL_631942\|2 | ···················· | ···················· | ····················· | ···················· | ···················· | ···················· |
| Q | 491 hCoV-19/USA/NY-NYCPHL-000475/2020\|EPI_ISL_631943\|2 | ···················· | ···················· | ····················· | ···················· | ···················· | ···················· |
| Q | 492 hCoV-19/USA/NY-NYCPHL-000477/2020\|EPI_ISL_631944\|2 | ···················· | ···················· | ····················· | ···················· | ···················· | ···················· |
| Q | 493 hCoV-19/USA/NY-NYCPHL-000478/2020\|EPI_ISL_631945\|2 | ···················· | ···················· | ····················· | ···················· | ···················· | ···················· |
| Q | 494 hCoV-19/USA/NY-NYCPHL-000479/2020\|EPI_ISL_631946\|2 | ···················· | ···················· | ····················· | ···················· | ···················· | ···················· |
| Q | 495 hCoV-19/USA/NY-NYCPHL-000483/2020\|EPI_ISL_631949\|2 | ···················· | ···················· | ····················· | ···················· | ···················· | ···················· |
| Q | 496 hCoV-19/USA/NY-NYCPHL-000485/2020\|EPI_ISL_631950\|2 | ···················· | ···················· | ····················· | ···················· | ···················· | ···················· |
| Q | 497 hCoV-19/USA/NY-NYCPHL-000486/2020\|EPI_ISL_631951\|2 | ···················· | ···················· | ····················· | ···················· | ···················· | ···················· |
| Q | 498 hCoV-19/USA/NY-NYCPHL-000487/2020\|EPI_ISL_631952\|2 | ···················· | ···················· | ····················· | ···················· | ···················· | ···················· |
| Q | 499 hCoV-19/USA/NY-NYCPHL-000490/2020\|EPI_ISL_631953\|2 | ···················· | ···················· | ····················· | ···················· | ···················· | ···················· |
| Q | 500 hCoV-19/USA/NY-NYCPHL-000491/2020\|EPI_ISL_631954\|2 | ···················· | ···················· | ····················· | ···················· | ···················· | ···················· |
| Q | 501 hCoV-19/USA/NY-NYCPHL-000492/2020\|EPI_ISL_631955\|2 | ···················· | ···················· | ····················· | ···················· | ···················· | ···················· |
| Q | 502 hCoV-19/USA/NY-NYCPHL-000494/2020\|EPI_ISL_631956\|2 | ···················· | ···················· | ····················· | ···················· | ···················· | ···················· |
| Q | 503 hCoV-19/USA/NY-NYCPHL-000495/2020\|EPI_ISL_631957\|2 | ···················· | ···················· | ····················· | ···················· | ···················· | ···················· |
| Q | 504 hCoV-19/USA/NY-NYCPHL-000496/2020\|EPI_ISL_631958\|2 | ···················· | ···················· | ····················· | ···················· | ···················· | ···················· |
| Q | 505 hCoV-19/USA/NY-NYCPHL-000498/2020\|EPI_ISL_631959\|2 | ···················· | ···················· | ····················· | ···················· | ···················· | ···················· |
| Q | 506 hCoV-19/USA/NY-NYCPHL-000499/2020\|EPI_ISL_631960\|2 | ···················· | ···················· | ····················· | ···················· | ···················· | ···················· |
| Q | 507 hCoV-19/USA/NY-NYCPHL-000500/2020\|EPI_ISL_631961\|2 | ···················· | ···················· | ····················· | ···················· | ···················· | ···················· |
| Q | 508 hCoV-19/USA/NY-NYCPHL-000501/2020\|EPI_ISL_631962\|2 | ···················· | ···················· | ····················· | ···················· | ···················· | ···················· |
| Q | 509 hCoV-19/USA/NY-NYCPHL-000524/2020\|EPI_ISL_631985\|2 | ···················· | ···················· | ····················· | ···················· | ···················· | ···················· |
| Q | 510 hCoV-19/USA/NY-NYCPHL-000525/2020\|EPI_ISL_631986\|2 | ···················· | ···················· | ····················· | ···················· | ···················· | ···················· |
| Q | 511 hCoV-19/USA/NY-NYCPHL-000526/2020\|EPI_ISL_631987\|2 | ···················· | ···················· | ····················· | ···················· | ···················· | ···················· |
| Q | 512 hCoV-19/USA/NY-NYCPHL-000527/2020\|EPI_ISL_631988\|2 | ···················· | ···················· | ····················· | ···················· | ···················· | ···················· |
| Q | 513 hCoV-19/Wales/PHWC-485BD1/2020\|EPI_ISL_628262\|2020 | ···················· | ···················· | ····················· | ···················· | ···················· | ···················· |
| Q | 514 hCoV-19/Wales/PHWC-485BFF/2020\|EPI_ISL_628263\|2020 | ···················· | ···················· | ····················· | ···················· | ·NN················· | ···················· |
| Q | 515 hCoV-19/Wales/PHWC-485C0B/2020\|EPI_ISL_628264\|2020 | ···················· | ···················· | ····················· | ···················· | ···················· | ···················· |
| Q | 516 hCoV-19/Wales/PHWC-485C1A/2020\|EPI_ISL_628265\|2020 | ···················· | ···················· | ····················· | ···················· | ···················· | ···················· |
| Q | 517 hCoV-19/Wales/PHWC-485C29/2020\|EPI_ISL_628266\|2020 | ···················· | ···················· | ····················· | ···················· | ···················· | ···················· |
| Q | 519 hCoV-19/Wales/PHWC-485C47/2020\|EPI_ISL_628268\|2020 | ···················· | ···················· | ····················· | ···················· | ···················· | ···················· |
| Q | 520 hCoV-19/Wales/PHWC-485C56/2020\|EPI_ISL_628269\|2020 | ···················· | ···················· | ····················· | ···················· | ···················· | ···················· |
| Q | 521 hCoV-19/England/MILK-AC84FF/2020\|EPI_ISL_629594\|20 | ···················· | ·A·················· | ····················· | ···················· | ···················· | ··A················· |
| Q | 522 hCoV-19/England/MILK-ACE81B/2020\|EPI_ISL_629595\|20 | ···················· | ···················· | ····················· | ···················· | ···················· | ···················· |
| Q | 523 hCoV-19/England/MILK-AC8019/2020\|EPI_ISL_629596\|20 | ···················· | ···················· | ····················· | ···················· | ···················· | ···················· |
| Q | 524 hCoV-19/England/MILK-AC894B/2020\|EPI_ISL_629598\|20 | ···················· | ···················· | ····················· | ···················· | ···················· | ···················· |
| Q | 525 hCoV-19/England/ALDP-A6779F/2020\|EPI_ISL_630577\|20 | ···················· | ···················· | ····················· | ···················· | ···················· | ···················· |
| Q | 526 hCoV-19/England/ALDP-A67058/2020\|EPI_ISL_630578\|20 | ···················· | ···················· | ····················· | ···················· | ···················· | ···················· |
| Q | 527 hCoV-19/England/ALDP-A68725/2020\|EPI_ISL_630579\|20 | ···················· | ···················· | ····················· | ···················· | ···················· | ···················· |
| Q | 528 hCoV-19/England/ALDP-A66B3A/2020\|EPI_ISL_630581\|20 | ···················· | ···················· | ····················· | ···················· | ···················· | ···················· |
| Q | 529 hCoV-19/England/ALDP-A67814/2020\|EPI_ISL_630582\|20 | ···················· | ···················· | ····················· | ···················· | ···················· | ···················· |
| Q | 530 hCoV-19/England/ALDP-A67331/2020\|EPI_ISL_630583\|20 | ···················· | ···················· | ····················· | ···················· | ···················· | ···················· |
| Q | 531 hCoV-19/England/ALDP-A67999/2020\|EPI_ISL_630586\|20 | ···················· | ···················· | ····················· | ···················· | ···················· | ···················· |
| Q | 532 hCoV-19/England/ALDP-A66A00/2020\|EPI_ISL_630587\|20 | ···················· | ···················· | ····················· | ···················· | ···················· | ···················· |
| Q | 533 hCoV-19/England/QEUH-AD3C08/2020\|EPI_ISL_634000\|20 | ···················· | ···················· | ····················· | ···················· | ···················· | ···················· |
| Q | 534 hCoV-19/England/QEUH-AD526E/2020\|EPI_ISL_634001\|20 | ···················· | ···················· | ····················· | ···················· | ···················· | ···················· |
| Q | 535 hCoV-19/England/QEUH-AD518F/2020\|EPI_ISL_634002\|20 | ···················· | ···················· | ····················· | ···················· | ···················· | ···················· |
| Q | 536 hCoV-19/England/QEUH-AD038E/2020\|EPI_ISL_634003\|20 | ···················· | ···················· | ····················· | ···················· | ···················· | ···················· |
| Q | 537 hCoV-19/England/QEUH-AD1FC5/2020\|EPI_ISL_634004\|20 | ···················· | ···················· | ····················· | ···················· | ···················· | ···················· |
| Q | 538 hCoV-19/England/QEUH-AD37DA/2020\|EPI_ISL_634005\|20 | ···················· | ···················· | ····················· | ···················· | ···················· | ···················· |
| Q | 539 hCoV-19/England/QEUH-AD3770/2020\|EPI_ISL_634006\|20 | ···················· | ···················· | ····················· | ···················· | ···················· | ···················· |
| Q | 540 hCoV-19/England/QEUH-AD40CF/2020\|EPI_ISL_634007\|20 | ···················· | ···················· | ····················· | ···················· | ···················· | ···················· |
| Q | 541 hCoV-19/England/QEUH-AD07BF/2020\|EPI_ISL_634008\|20 | ···················· | ···················· | ····················· | ···················· | ···················· | ···················· |
| Q | 542 hCoV-19/England/QEUH-AD2911/2020\|EPI_ISL_634009\|20 | ···················· | ···················· | ····················· | ···················· | ···················· | ···················· |
| Q | 543 hCoV-19/England/QEUH-AD3813/2020\|EPI_ISL_634010\|20 | ···················· | ·A·················· | ····················· | ···················· | ···················· | ···················· |
| Q | 544 hCoV-19/England/QEUH-AD0588/2020\|EPI_ISL_634011\|20 | ···················· | ···················· | ····················· | ···················· | ···················· | ···················· |
| Q | 545 hCoV-19/England/QEUH-AD3725/2020\|EPI_ISL_634012\|20 | ···················· | ···················· | ····················· | ···················· | ···················· | ···················· |
| Q | 546 hCoV-19/England/QEUH-AD297B/2020\|EPI_ISL_634013\|20 | ···················· | ···················· | ····················· | ···················· | ···················· | ···················· |
| Q | 547 hCoV-19/England/QEUH-AD2085/2020\|EPI_ISL_634014\|20 | ···················· | ···················· | ····················· | ···················· | ···················· | ···················· |
| Q | 548 hCoV-19/England/QEUH-AD30A2/2020\|EPI_ISL_634015\|20 | ···················· | ···················· | ····················· | ···················· | ···················· | ···················· |
| Q | 549 hCoV-19/England/QEUH-AD48C7/2020\|EPI_ISL_634016\|20 | ···················· | ···················· | ····················· | ···················· | ···················· | ···················· |
| Q | 550 hCoV-19/England/QEUH-AD0C65/2020\|EPI_ISL_634017\|20 | ···················· | ···················· | ····················· | ···················· | ···················· | ···················· |
| Q | 551 hCoV-19/England/QEUH-AD236E/2020\|EPI_ISL_634019\|20 | ···················· | ···················· | ····················· | ···················· | ···················· | ···················· |
| Q | 552 hCoV-19/England/QEUH-AD30B1/2020\|EPI_ISL_634031\|20 | ···················· | ···················· | ····················· | ···················· | ···················· | ···················· |
| Q | 553 hCoV-19/England/QEUH-AD3CBD/2020\|EPI_ISL_634032\|20 | ···················· | ·A·················· | ····················· | ···················· | ···················· | ··A················· |
| Q | 554 hCoV-19/England/QEUH-AD3BFC/2020\|EPI_ISL_634033\|20 | ···················· | ···················· | ····················· | ···················· | ···················· | ···················· |
| Q | 555 hCoV-19/England/QEUH-AD66BC/2020\|EPI_ISL_634040\|20 | ···················· | ···················· | ····················· | ···················· | ···················· | ···················· |
| Q | 556 hCoV-19/England/QEUH-AD5134/2020\|EPI_ISL_634042\|20 | ···················· | ···················· | ····················· | ···················· | ···················· | ···················· |

Q 557 hCoV-19/England/QEUH-AD33D6/2020|EPI_ISL_634044|20 ···················· ···················· ····················· ···················· ····K··············· ····················
Q 558 hCoV-19/England/QEUH-AD493D/2020|EPI_ISL_634045|20 ···················· ···················· ····················· ···················· ···················· ····················
Q 559 hCoV-19/England/QEUH-AD3CAE/2020|EPI_ISL_634046|20 ···················· ···················· ····················· ···················· ···················· ····················
Q 560 hCoV-19/England/QEUH-AD4900/2020|EPI_ISL_634047|20 ···················· ···················· ····················· ···················· ···················· ····················
Q 561 hCoV-19/England/QEUH-AD50BF/2020|EPI_ISL_634048|20 ···················· ···················· ····················· ···················· ···················· ····················
Q 562 hCoV-19/England/QEUH-AD0CCF/2020|EPI_ISL_634049|20 ···················· ···················· ····················· ···················· ···················· ····················
Q 563 hCoV-19/England/QEUH-AD51F8/2020|EPI_ISL_634050|20 ···················· ·A·················· ····················· ···················· ···················· ···A················
Q 564 hCoV-19/England/QEUH-AD23B9/2020|EPI_ISL_634051|20 ···················· ···················· ····················· ···················· ···················· ····················
Q 565 hCoV-19/England/QEUH-AD5231/2020|EPI_ISL_634052|20 ···················· ···················· ····················· ···················· ···················· ····················
Q 566 hCoV-19/England/QEUH-AD4A58/2020|EPI_ISL_634054|20 ···················· ···················· ····················· ···················· ···················· ····················
Q 567 hCoV-19/England/QEUH-AD3039/2020|EPI_ISL_634055|20 ···················· ···················· ····················· ···················· ···················· ····················
Q 568 hCoV-19/England/QEUH-AD23AA/2020|EPI_ISL_634056|20 ···················· ···················· ····················· ···················· ···················· ····················
Q 569 hCoV-19/England/QEUH-AD50FB/2020|EPI_ISL_634057|20 ···················· ···················· ····················· ···················· ···················· ····················
Q 570 hCoV-19/England/QEUH-AD0D71/2020|EPI_ISL_634058|20 ···················· ···················· ····················· ···················· ···················· ····················
Q 571 hCoV-19/England/QEUH-AD2CBE/2020|EPI_ISL_634059|20 ···················· ···················· ····················· ···················· ···················· ····················
Q 572 hCoV-19/England/QEUH-AD2BA2/2020|EPI_ISL_634060|20 ···················· ···················· ····················· ···················· ···················· ····················
Q 573 hCoV-19/England/QEUH-AD3C80/2020|EPI_ISL_634061|20 ···················· ···················· ····················· ···················· ···················· ····················
Q 574 hCoV-19/England/QEUH-AD237D/2020|EPI_ISL_634062|20 ···················· ···················· ····················· ···················· ···················· ····················
Q 575 hCoV-19/England/QEUH-AD3C9F/2020|EPI_ISL_634064|20 ···················· ···················· ····················· ···················· ···················· ····················
Q 576 hCoV-19/England/QEUH-AD3CF9/2020|EPI_ISL_634065|20 ···················· ···················· ····················· ···················· ···················· ····················
Q 577 hCoV-19/England/QEUH-AD2C54/2020|EPI_ISL_634066|20 ···················· ···················· ····················· ···················· ···················· ····················
Q 578 hCoV-19/England/QEUH-AD3C53/2020|EPI_ISL_634067|20 ···················· ···················· ····················· ···················· ···················· ····················
Q 579 hCoV-19/England/QEUH-AD3707/2020|EPI_ISL_634068|20 ···················· ···················· ····················· ···················· ···················· ····················
Q 580 hCoV-19/England/QEUH-AD5213/2020|EPI_ISL_634069|20 ···················· ···················· ····················· ···················· ···················· ····················
Q 581 hCoV-19/England/QEUH-AD0263/2020|EPI_ISL_634070|20 ···················· ···················· ····················· ···················· ···················· ····················
Q 582 hCoV-19/England/QEUH-AD49A6/2020|EPI_ISL_634071|20 ···················· ···················· ····················· ···················· ···················· ····················
Q 583 hCoV-19/England/QEUH-AD5222/2020|EPI_ISL_634073|20 ···················· ···················· ····················· ···················· ···················· ····················
Q 584 hCoV-19/Wales/QEUH-AD36BF/2020|EPI_ISL_634074|2020 ···················· ···················· ····················· ···················· ···················· ····················
Q 585 hCoV-19/England/QEUH-AD527D/2020|EPI_ISL_634075|20 ···················· ···················· ····················· ···················· ···················· ····················
Q 586 hCoV-19/England/QEUH-AD4135/2020|EPI_ISL_634076|20 ···················· ···················· ····················· ···················· ···················· ····················
Q 587 hCoV-19/England/QEUH-AD2C36/2020|EPI_ISL_634077|20 ···················· ···················· ····················· ···················· ···················· ····················
Q 588 hCoV-19/England/QEUH-AD2B39/2020|EPI_ISL_634078|20 ···················· ·A·················· ····················· ···················· ···················· ···A················
Q 589 hCoV-19/England/QEUH-AD488B/2020|EPI_ISL_634079|20 ···················· ···················· ····················· ···················· ···················· ····················
Q 590 hCoV-19/England/QEUH-AD6643/2020|EPI_ISL_634100|20 ···················· ···················· ····················· ···················· ···················· ····················
Q 591 hCoV-19/England/QEUH-AD2067/2020|EPI_ISL_634101|20 ···················· ···················· ····················· ···················· ···················· ····················
Q 592 hCoV-19/England/QEUH-AD20C1/2020|EPI_ISL_634102|20 ···················· ···················· ····················· ···················· ···················· ····················
Q 593 hCoV-19/England/QEUH-AD0412/2020|EPI_ISL_634103|20 ···················· ···················· ····················· ···················· ···················· ····················
Q 594 hCoV-19/England/QEUH-AD0D62/2020|EPI_ISL_634104|20 ···················· ···················· ····················· ···················· ···················· ····················
Q 595 hCoV-19/England/QEUH-AD4988/2020|EPI_ISL_634105|20 ···················· ···················· ····················· ···················· ···················· ····················
Q 596 hCoV-19/England/QEUH-AD37AD/2020|EPI_ISL_634106|20 ···················· ···················· ····················· ···················· ···················· ····················
Q 597 hCoV-19/England/QEUH-AD2BB1/2020|EPI_ISL_634110|20 ···················· ···················· ····················· ···················· ···················· ····················
Q 598 hCoV-19/England/QEUH-AD2401/2020|EPI_ISL_634111|20 ···················· ·A·················· ····················· ···················· ···················· ···A················
Q 599 hCoV-19/England/QEUH-AD246B/2020|EPI_ISL_634112|20 ···················· ···················· ····················· ···················· ···················· ····················
Q 600 hCoV-19/England/QEUH-AD3CDB/2020|EPI_ISL_634114|20 ···················· ···················· ····················· ···················· ···················· ····················
Q 601 hCoV-19/England/QEUH-AD4A0D/2020|EPI_ISL_634115|20 ···················· ···················· ····················· ···················· ···················· ····················
Q 602 hCoV-19/England/QEUH-AD0C92/2020|EPI_ISL_634116|20 ···················· ···················· ····················· ···················· ···················· ····················
Q 603 hCoV-19/England/QEUH-AD48A9/2020|EPI_ISL_634117|20 ···················· ···················· ····················· ···················· ···················· ····················
Q 604 hCoV-19/England/QEUH-AD2BDF/2020|EPI_ISL_634118|20 ···················· ···················· ····················· ···················· ···················· ····················
Q 605 hCoV-19/USA/CA-ALSR-2064/2020|EPI_ISL_635329|2020- ···················· ·A·················· ····················· ···················· ···················· ····················
Q 606 hCoV-19/USA/CA-ALSR-2065/2020|EPI_ISL_635330|2020- ···················· ·A·················· ····················· ···················· ···················· ····················
Q 607 hCoV-19/USA/CA-ALSR-2066/2020|EPI_ISL_635331|2020- ···················· ·A·················· ····················· ···················· ···················· ····················
Q 608 hCoV-19/USA/CA-ALSR-2067/2020|EPI_ISL_635332|2020- ···················· ·A·················· ····················· ···················· ···················· ····················
Q 609 hCoV-19/USA/CA-ALSR-2069/2020|EPI_ISL_635333|2020- ···················· ·A·················· ····················· ···················· ···················· ····················
Q 610 hCoV-19/USA/CA-ALSR-2070/2020|EPI_ISL_635334|2020- ···················· ·A·················· ····················· ···················· ···················· ····················
Q 611 hCoV-19/USA/CA-ALSR-2073/2020|EPI_ISL_635335|2020- ···················· ·A·················· ····················· ···················· ···················· ····················
Q 612 hCoV-19/USA/CA-ALSR-2076/2020|EPI_ISL_635336|2020- ···················· ·A·················· ····················· ···················· ···················· ····················
Q 613 hCoV-19/USA/CA-ALSR-2079/2020|EPI_ISL_635337|2020- ···················· ·A·················· ····················· ···················· ···················· ····················
Q 614 hCoV-19/USA/CA-ALSR-2081/2020|EPI_ISL_635338|2020- ···················· ·A·················· ····················· ···················· ···················· ····················
Q 615 hCoV-19/USA/CA-ALSR-2085/2020|EPI_ISL_635340|2020- ···················· ·A·················· ····················· ···················· ···················· ····················
Q 616 hCoV-19/USA/CA-ALSR-2086/2020|EPI_ISL_635341|2020- ···················· ·A·················· ····················· ···················· ···················· ····················
Q 617 hCoV-19/USA/CA-ALSR-2089/2020|EPI_ISL_635342|2020- ···················· ·A·················· ····················· ···················· ···················· ····················
Q 618 hCoV-19/USA/CA-ALSR-2092/2020|EPI_ISL_635343|2020- ···················· ·A·················· ····················· ···················· ···················· ····················
Q 619 hCoV-19/USA/CA-ALSR-2093/2020|EPI_ISL_635344|2020- ···················· ·A·················· ····················· ···················· ···················· ····················
Q 620 hCoV-19/USA/CA-ALSR-2094/2020|EPI_ISL_635345|2020- ···················· ···················· ····················· ···················· ···················· ····················
Q 621 hCoV-19/USA/CA-ALSR-2096/2020|EPI_ISL_635346|2020- ···················· ·A·················· ····················· ···················· ···················· ····················
Q 622 hCoV-19/USA/CA-ALSR-2099/2020|EPI_ISL_635347|2020- ···················· ·A·················· ····················· ···················· ···················· ····················
Q 623 hCoV-19/USA/CA-ALSR-2105/2020|EPI_ISL_635348|2020- ···················· ···················· ····················· ···················· ···················· ····················
Q 624 hCoV-19/USA/CA-ALSR-2106/2020|EPI_ISL_635349|2020- ···················· ·A·················· ····················· ···················· ···················· ····················
Q 625 hCoV-19/USA/CA-ALSR-2133/2020|EPI_ISL_635361|2020- ···················· ·A·················· ····················· ···················· ···················· ····················
Q 626 hCoV-19/USA/CA-ALSR-2158/2020|EPI_ISL_635363|2020- ···················· ·A·················· ····················· ···················· ···················· ····················
Q 627 hCoV-19/USA/CA-ALSR-2160/2020|EPI_ISL_635364|2020- ···················· ·A·················· ····················· ···················· ···················· ····················
Q 628 hCoV-19/USA/CA-ALSR-2169/2020|EPI_ISL_635370|2020- ···················· ·A·················· ····················· ···················· ···················· ····················
Q 629 hCoV-19/USA/CA-ALSR-2170/2020|EPI_ISL_635371|2020- ···················· ···················· ····················· ···················· ···················· ····················
Q 630 hCoV-19/USA/CA-ALSR-2171/2020|EPI_ISL_635372|2020- ···················· ···················· ····················· ···················· ···················· ····················
Q 631 hCoV-19/USA/CA-ALSR-2172/2020|EPI_ISL_635373|2020- ···················· ···················· ····················· ···················· ···················· ····················
Q 632 hCoV-19/USA/CA-ALSR-2174/2020|EPI_ISL_635374|2020- ···················· ·A·················· ····················· ···················· ···················· ····················
Q 633 hCoV-19/USA/CA-ALSR-2175/2020|EPI_ISL_635375|2020- ···················· ·A·················· ····················· ···················· ···················· ····················
Q 634 hCoV-19/USA/CA-ALSR-2179/2020|EPI_ISL_635376|2020- ···················· ···················· ····················· ···················· ···················· ····················
Q 635 hCoV-19/USA/CA-ALSR-2180/2020|EPI_ISL_635377|2020- ···················· ···················· ····················· ···················· ···················· ····················
Q 636 hCoV-19/USA/CA-ALSR-2183/2020|EPI_ISL_635378|2020- ···················· ···················· ····················· ···················· ···················· ····················

```
Q    637 hCoV-19/USA/CA-ALSR-2185/2020|EPI_ISL_635379|2020-   ····················  ·A··················  ····················  ····················  ····················  ····················
Q    638 hCoV-19/USA/CA-ALSR-2188/2020|EPI_ISL_635380|2020-   ····················  ····················  ····················  ····················  ····················  ····················
Q    639 hCoV-19/USA/CA-ALSR-2190/2020|EPI_ISL_635381|2020-   ····················  ····················  ····················  ····················  ····················  ····················
Q    640 hCoV-19/USA/CA-ALSR-2192/2020|EPI_ISL_635382|2020-   ····················  ·A··················  ····················  ····················  ····················  ····················
Q    641 hCoV-19/USA/CA-ALSR-2195/2020|EPI_ISL_635383|2020-   ····················  ····················  ····················  ····················  ····················  ····················
Q    642 hCoV-19/USA/CA-ALSR-2201/2020|EPI_ISL_635385|2020-   ····················  ····················  ····················  ····················  ····················  ····················
Q    643 hCoV-19/USA/CA-ALSR-2202/2020|EPI_ISL_635386|2020-   ····················  ·A··················  ····················  ····················  ····················  ····················
Q    644 hCoV-19/USA/CA-ALSR-2207/2020|EPI_ISL_635387|2020-   ····················  ·A··················  ····················  ····················  ····················  ····················
Q    645 hCoV-19/USA/CA-ALSR-2213/2020|EPI_ISL_635388|2020-   ····················  ····················  ····················  ····················  ····················  ····················
Q    646 hCoV-19/USA/CA-ALSR-2216/2020|EPI_ISL_635389|2020-   ····················  ·A··················  ····················  ····················  ····················  ····················
Q    647 hCoV-19/USA/CA-ALSR-2218/2020|EPI_ISL_635390|2020-   ····················  ·A··················  ····················  ····················  ····················  ····················
Q    648 hCoV-19/USA/CA-ALSR-2219/2020|EPI_ISL_635391|2020-   ····················  ·A··················  ····················  ····················  ····················  ····················
Q    649 hCoV-19/USA/CA-ALSR-2223/2020|EPI_ISL_635392|2020-   ····················  ·A··················  ····················  ····················  ····················  ····················
Q    650 hCoV-19/USA/CA-ALSR-2225/2020|EPI_ISL_635393|2020-   ····················  ····················  ····················  ····················  ····················  ····················
Q    651 hCoV-19/USA/CA-ALSR-2230/2020|EPI_ISL_635394|2020-   ····················  ····················  ····················  ····················  ····················  ····················
Q    652 hCoV-19/USA/CA-ALSR-2231/2020|EPI_ISL_635395|2020-   ········T···········  ·A··················  ····················  ····················  ····················  ····················
Q    653 hCoV-19/USA/CA-ALSR-2232/2020|EPI_ISL_635396|2020-   ····················  ····················  ····················  ····················  ····················  ····················
Q    654 hCoV-19/USA/CA-ALSR-2234/2020|EPI_ISL_635397|2020-   ····················  ····················  ····················  ····················  ····················  ····················
Q    655 hCoV-19/USA/CA-ALSR-2236/2020|EPI_ISL_635398|2020-   ····················  ·A··················  ····················  ····················  ····················  ····················
Q    656 hCoV-19/USA/CA-ALSR-2237/2020|EPI_ISL_635399|2020-   ····················  ·A··················  ····················  ····················  ····················  ····················
Q    657 hCoV-19/USA/CA-ALSR-2268/2020|EPI_ISL_635401|2020-   ····················  ·A··················  ····················  ····················  ····················  ····················
Q    658 hCoV-19/USA/CA-ALSR-2269/2020|EPI_ISL_635402|2020-   ····················  ·A··················  ····················  ····················  ····················  ····················
Q    659 hCoV-19/USA/CA-ALSR-2270/2020|EPI_ISL_635403|2020-   ····················  ····················  ····················  ····················  ····················  ····················
Q    660 hCoV-19/USA/CA-ALSR-2271/2020|EPI_ISL_635404|2020-   ····················  ·A··················  ····················  ····················  ····················  ····················
Q    661 hCoV-19/USA/CA-ALSR-2276/2020|EPI_ISL_635405|2020-   ····················  ····················  ····················  ····················  ····················  ····················
Q    662 hCoV-19/USA/CA-ALSR-2279/2020|EPI_ISL_635406|2020-   ····················  ·A··················  ····················  ····················  ····················  ····················
Q    663 hCoV-19/USA/CA-ALSR-2280/2020|EPI_ISL_635407|2020-   ····················  ·A··················  ····················  ····················  ····················  ····················
Q    664 hCoV-19/USA/CA-ALSR-2287/2020|EPI_ISL_635412|2020-   ····················  ·A··················  ····················  ····················  ····················  ····················
Q    665 hCoV-19/USA/CA-ALSR-2289/2020|EPI_ISL_635413|2020-   ····················  ····················  ····················  ····················  ····················  ····················
Q    666 hCoV-19/USA/CA-ALSR-2290/2020|EPI_ISL_635414|2020-   ····················  ·A··················  ····················  ····················  ····················  ····················
Q    667 hCoV-19/USA/CA-ALSR-2292/2020|EPI_ISL_635416|2020-   ····················  ····················  ····················  ····················  ····················  ····················
Q    668 hCoV-19/USA/CA-ALSR-2294/2020|EPI_ISL_635417|2020-   ····················  ····················  ····················  ····················  ····················  ····················
Q    669 hCoV-19/USA/CA-ALSR-2297/2020|EPI_ISL_635418|2020-   ····················  ·A··················  ····················  ····················  ····················  ····················
Q    670 hCoV-19/USA/CA-ALSR-2299/2020|EPI_ISL_635420|2020-   ····················  ·A··················  ····················  ····················  ····················  ····················
Q    671 hCoV-19/USA/CA-ALSR-2302/2020|EPI_ISL_635421|2020-   ····················  ·A··················  ····················  ····················  ····················  ····················
Q    672 hCoV-19/USA/CA-ALSR-2306/2020|EPI_ISL_635422|2020-   ····················  ·A··················  ····················  ····················  ····················  ····················
Q    673 hCoV-19/USA/CA-ALSR-2308/2020|EPI_ISL_635423|2020-   ····················  ·A··················  ····················  ····················  ····················  ····················
Q    674 hCoV-19/USA/CA-ALSR-2310/2020|EPI_ISL_635424|2020-   ···········T········  ····················  ····················  ····················  ····················  ····················
Q    675 hCoV-19/USA/CA-ALSR-2314/2020|EPI_ISL_635425|2020-   ···········T········  ····················  ····················  ····················  ····················  ····················
Q    676 hCoV-19/USA/CA-ALSR-2317/2020|EPI_ISL_635427|2020-   ····················  ·A··················  ····················  ····················  ····················  ····················
Q    677 hCoV-19/USA/CA-ALSR-2318/2020|EPI_ISL_635428|2020-   ····················  ·A··················  ····················  ····················  ····················  ····················
Q    678 hCoV-19/USA/CA-ALSR-2320/2020|EPI_ISL_635429|2020-   ····················  ····················  ····················  ····················  ····················  ····················
Q    679 hCoV-19/USA/CA-ALSR-2321/2020|EPI_ISL_635430|2020-   ····················  ·A··················  ····················  ····················  ····················  ····················
Q    680 hCoV-19/USA/CA-ALSR-2327/2020|EPI_ISL_635432|2020-   ····················  ····················  ····················  ····················  ····················  ····················
Q    681 hCoV-19/USA/CA-ALSR-2332/2020|EPI_ISL_635433|2020-   ····················  ·A··················  ····················  ····················  ····················  ····················
Q    682 hCoV-19/USA/CA-ALSR-2335/2020|EPI_ISL_635435|2020-   ····················  ·A·······A··········  ····················  ····················  ····················  ····················
Q    683 hCoV-19/USA/CA-ALSR-2337/2020|EPI_ISL_635436|2020-   ····················  ····················  ····················  ····················  ····················  ····················
Q    684 hCoV-19/USA/CA-ALSR-2338/2020|EPI_ISL_635437|2020-   ····················  ·A··················  ····················  ····················  ····················  ····················
Q    685 hCoV-19/USA/CA-ALSR-2344/2020|EPI_ISL_635438|2020-   ····················  ····················  ····················  ····················  ····················  ····················
Q    686 hCoV-19/USA/CA-ALSR-2346/2020|EPI_ISL_635439|2020-   ····················  ·A··················  ····················  ····················  ····················  ····················
Q    687 hCoV-19/USA/CA-ALSR-2353/2020|EPI_ISL_635440|2020-   ····················  ·A··················  ····················  ····················  ····················  ····················
Q    688 hCoV-19/USA/CA-ALSR-2359/2020|EPI_ISL_635441|2020-   ····················  ····················  ····················  ····················  ····················  ····················
Q    689 hCoV-19/USA/CA-ALSR-2362/2020|EPI_ISL_635442|2020-   ····················  ·A··················  ····················  ····················  ····················  ····················
Q    690 hCoV-19/USA/CA-ALSR-2367/2020|EPI_ISL_635444|2020-   ····················  ·A··················  ····················  ····················  ····················  ····················
Q    691 hCoV-19/USA/CA-ALSR-2368/2020|EPI_ISL_635445|2020-   ····················  ····················  ····················  ····················  ····················  ····················
Q    692 hCoV-19/USA/CA-ALSR-2369/2020|EPI_ISL_635446|2020-   ····················  ·A··················  ····················  ····················  ····················  ····················
Q    693 hCoV-19/USA/CA-ALSR-2372/2020|EPI_ISL_635447|2020-   ····················  ·A··················  ····················  ····················  ····················  ····················
Q    694 hCoV-19/USA/CA-ALSR-2377/2020|EPI_ISL_635448|2020-   ····················  ·A··················  ····················  ····················  ····················  ····················
Q    695 hCoV-19/USA/CA-ALSR-2378/2020|EPI_ISL_635449|2020-   ····················  ····················  ····················  ····················  ····················  ····················
Q    708 hCoV-19/Belgium/ULG-10700/2020|EPI_ISL_636672|2020   ····················  ····················  ····················  ····················  ····················  ····················
Q    711 hCoV-19/Belgium/ULG-10703/2020|EPI_ISL_636675|2020   ····················  ····················  ····················  ····················  ····················  ····················
Q    715 hCoV-19/England/C200028993/2020|EPI_ISL_636696|202   ····················  ····················  ····················  ····················  ····················  ····················
Q    716 hCoV-19/England/C200028994/2020|EPI_ISL_636697|202   ····················  ····················  ····················  ····················  ····················  ····················
Q    717 hCoV-19/England/C200029067/2020|EPI_ISL_636700|202   ····················  ····················  ····················  ····················  ····················  ····················
Q    718 hCoV-19/England/C200029071/2020|EPI_ISL_636701|202   ····················  ····················  ····················  ····················  ····················  ····················
Q    719 hCoV-19/England/C200029186/2020|EPI_ISL_636702|202   ····················  ····················  ····················  ····················  ····················  ····················
Q    720 hCoV-19/England/C200029520/2020|EPI_ISL_636703|202   ····················  ····················  ····················  ····················  ····················  ····················
Q    721 hCoV-19/England/C200032612/2020|EPI_ISL_636705|202   ····················  ····················  ····················  ····················  ······K·············  ····················
Q    722 hCoV-19/England/C200032707/2020|EPI_ISL_636706|202   ····················  ····················  ····················  ····················  ····················  ····················
Q    723 hCoV-19/England/C200032709/2020|EPI_ISL_636707|202   ····················  ····················  ····················  ····················  ····················  ····················
Q    724 hCoV-19/England/C200032712/2020|EPI_ISL_636708|202   ····················  ····················  ····················  ····················  ····················  ····················
Q    725 hCoV-19/England/C200032718/2020|EPI_ISL_636709|202   ····················  ····················  ····················  ····················  ····················  ····················
Q    726 hCoV-19/England/C200032834/2020|EPI_ISL_636710|202   ····················  ····················  ····················  ····················  ····················  ····················
Q    727 hCoV-19/England/C200032839/2020|EPI_ISL_636711|202   ····················  ····················  ····················  ····················  ····················  ····················
Q    728 hCoV-19/England/C200032845/2020|EPI_ISL_636712|202   ····················  ····················  ····················  ····················  ····················  ····················
Q    729 hCoV-19/England/C200032894/2020|EPI_ISL_636714|202   ····················  ····················  ····················  ····················  ····················  ····················
Q    730 hCoV-19/England/C200032897/2020|EPI_ISL_636715|202   ····················  ····················  ····················  ····················  ····················  ····················
Q    731 hCoV-19/England/C200035171/2020|EPI_ISL_636716|202   ····················  ····················  ····················  ····················  ····················  ····················
Q    732 hCoV-19/England/C200035466/2020|EPI_ISL_636718|202   ····················  ····················  ····················  ····················  ····················  ····················
Q    733 hCoV-19/England/C200036080/2020|EPI_ISL_636719|202   ····················  ····················  ····················  ····················  ····················  ····················
```

| | | | | | | | | |
|---|---|---|---|---|---|---|---|---|
| Q | 734 hCoV-19/England/C200036082/2020\|EPI_ISL_636720\|202 | ···················· | ···················· | ···················· | ···················· | ···················· | ···················· | |
| Q | 735 hCoV-19/England/C200036087/2020\|EPI_ISL_636721\|202 | ···················· | ···················· | ···················· | ···················· | ···················· | ···················· | |
| Q | 736 hCoV-19/England/C200036211/2020\|EPI_ISL_636722\|202 | ···················· | ···················· | ···················· | ···················· | ···················· | ···················· | |
| Q | 737 hCoV-19/England/C200036256/2020\|EPI_ISL_636723\|202 | ···················· | ···················· | ···················· | ···················· | ···················· | ···················· | |
| Q | 738 hCoV-19/England/C200036265/2020\|EPI_ISL_636724\|202 | ···················· | ···················· | ···················· | ···················· | ···················· | ···················· | |
| Q | 739 hCoV-19/England/C200037134/2020\|EPI_ISL_636728\|202 | ···················· | ···················· | ···················· | ···················· | ···················· | ···················· | |
| Q | 740 hCoV-19/England/C200037197/2020\|EPI_ISL_636729\|202 | ···················· | ···················· | ···················· | ···················· | ···················· | ···················· | |
| Q | 741 hCoV-19/England/C200037229/2020\|EPI_ISL_636730\|202 | ···················· | ···················· | ···················· | ···················· | ···················· | ···················· | |
| Q | 742 hCoV-19/England/C200040575/2020\|EPI_ISL_636731\|202 | ···················· | ···················· | ···················· | ···················· | ···················· | ···················· | |
| Q | 743 hCoV-19/England/C200040625/2020\|EPI_ISL_636732\|202 | ···················· | ···················· | ···················· | ···················· | ···················· | ···················· | |
| Q | 744 hCoV-19/England/C200040705/2020\|EPI_ISL_636733\|202 | ···················· | ···················· | ···················· | ···················· | ···················· | ···················· | |
| Q | 745 hCoV-19/England/204620001/2020\|EPI_ISL_636734\|2020 | ···················· | ···················· | ···················· | ···················· | ···················· | ···················· | |
| Q | 746 hCoV-19/Brazil/RJ-06020/2020\|EPI_ISL_636737\|2020-0 | ···················· | ···················· | ···················· | ···················· | ···················· | ···················· | |
| Q | 747 hCoV-19/India/DL-57012-CSIR-IGIB/2020\|EPI_ISL_6367 | ···················· | ···················· | ···················· | ···················· | ···················· | ···················· | |
| Q | 748 hCoV-19/India/UT-67339-CSIR-IGIB/2020\|EPI_ISL_6367 | ···················· | ·A·················· | ···················· | ···················· | ···················· | ··A················· | |
| Q | 749 hCoV-19/India/UT-67341-CSIR-IGIB/2020\|EPI_ISL_6367 | ···················· | ·A·················· | ···················· | ···················· | ···················· | ··A················· | |
| Q | 750 hCoV-19/India/UT-67340-CSIR-IGIB/2020\|EPI_ISL_6367 | ···················· | ·A·················· | ···················· | ···················· | ···················· | ··A················· | |
| Q | 751 hCoV-19/India/UT-67330-CSIR-IGIB/2020\|EPI_ISL_6367 | ···················· | ·A·················· | ···················· | ···················· | ···················· | ··A················· | |
| Q | 752 hCoV-19/India/UT-50438-CSIR-IGIB/2020\|EPI_ISL_6367 | ···················· | ·A·················· | ···················· | ···················· | ···················· | ··A················· | |
| Q | 753 hCoV-19/India/UT-67177-CSIR-IGIB/2020\|EPI_ISL_6367 | ···················· | ·A·················· | ···················· | ···················· | ···················· | ··A················· | |
| Q | 754 hCoV-19/India/DL-59946-CSIR-IGIB/2020\|EPI_ISL_6367 | ···················· | ·A·················· | ···················· | ···················· | ···················· | ··A················· | |
| Q | 755 hCoV-19/India/UT-67134-CSIR-IGIB/2020\|EPI_ISL_6367 | ···················· | ·A·················· | ···················· | ···················· | ···················· | ··A················· | |
| Q | 756 hCoV-19/India/DL-69833-CSIR-IGIB/2020\|EPI_ISL_6367 | ···················· | ···················· | ···················· | ···················· | ···················· | ···················· | |
| Q | 757 hCoV-19/India/DL-69831-CSIR-IGIB/2020\|EPI_ISL_6367 | ···················· | ·A·················· | ···················· | ···················· | ···················· | ··A················· | |
| Q | 758 hCoV-19/India/UT-67165-CSIR-IGIB/2020\|EPI_ISL_6367 | ···················· | ·A·················· | ···················· | ···················· | ···················· | ··A················· | |
| Q | 759 hCoV-19/India/DL-69826-CSIR-IGIB/2020\|EPI_ISL_6367 | ···················· | ·A·················· | ···················· | ···················· | ···················· | ··A················· | |
| Q | 760 hCoV-19/India/UT-50464-CSIR-IGIB/2020\|EPI_ISL_6367 | ···················· | ···················· | ··················C· | ···················· | ···················· | ···················· | 1 |
| Q | 761 hCoV-19/India/DL-69787-CSIR-IGIB/2020\|EPI_ISL_6367 | ···················· | ·A·················· | ···················· | ···················· | ···················· | ··A················· | |
| Q | 762 hCoV-19/India/DL-69794-CSIR-IGIB/2020\|EPI_ISL_6367 | ···················· | ···················· | ···················· | ···················· | ···················· | ···················· | |
| Q | 763 hCoV-19/India/DL-69813-CSIR-IGIB/2020\|EPI_ISL_6367 | ···················· | ·A·················· | ···················· | ···················· | ···················· | ··A················· | |
| Q | 764 hCoV-19/India/DL-61026-CSIR-IGIB/2020\|EPI_ISL_6367 | ···················· | ·A·················· | ···················· | ···················· | ···················· | ··A················· | |
| Q | 765 hCoV-19/India/DL-50480-CSIR-IGIB/2020\|EPI_ISL_6367 | ···················· | ·A·················· | ···················· | ···················· | ···················· | ··A················· | |
| Q | 766 hCoV-19/India/DL-60920-CSIR-IGIB/2020\|EPI_ISL_6367 | ···················· | ·A·················· | ···················· | ···················· | ···················· | ··A················· | |
| Q | 767 hCoV-19/India/DL-62368-CSIR-IGIB/2020\|EPI_ISL_6367 | ···················· | ·A·················· | ···················· | ···················· | ···················· | ··A················· | |
| Q | 768 hCoV-19/India/DL-69775-CSIR-IGIB/2020\|EPI_ISL_6367 | ···················· | ·A·················· | ···················· | ···················· | ···················· | ··A················· | |
| Q | 769 hCoV-19/Wales/PHWC-48561C/2020\|EPI_ISL_628200\|2020 | ···················· | ···················· | ···················· | ···················· | ···················· | ···················· | |
| Q | 770 hCoV-19/Wales/PHWC-485649/2020\|EPI_ISL_628201\|2020 | ···················· | ···················· | ···················· | ···················· | ···················· | ···················· | |
| Q | 771 hCoV-19/Wales/PHWC-485658/2020\|EPI_ISL_628202\|2020 | ···················· | ···················· | ···················· | ···················· | ···················· | ···················· | |
| Q | 772 hCoV-19/Wales/PHWC-485667/2020\|EPI_ISL_628203\|2020 | ···················· | ···················· | ···················· | ···················· | ···················· | ···················· | |
| Q | 773 hCoV-19/Wales/PHWC-485676/2020\|EPI_ISL_628204\|2020 | ···················· | ···················· | ···················· | ···················· | ···················· | ···················· | |
| Q | 775 hCoV-19/Wales/PHWC-4856B2/2020\|EPI_ISL_628206\|2020 | ···················· | ···················· | ···················· | ···················· | ···················· | ···················· | |
| Q | 776 hCoV-19/Wales/PHWC-4856D0/2020\|EPI_ISL_628207\|2020 | ···················· | ···················· | ···················· | ···················· | ···················· | ···················· | |
| Q | 777 hCoV-19/Wales/PHWC-4856EF/2020\|EPI_ISL_628208\|2020 | ···················· | ···················· | ···················· | ···················· | ···················· | ···················· | |
| Q | 778 hCoV-19/Wales/PHWC-4856FE/2020\|EPI_ISL_628209\|2020 | ···················· | ···················· | ···················· | ···················· | ···················· | ···················· | |
| Q | 779 hCoV-19/Wales/PHWC-48570A/2020\|EPI_ISL_628210\|2020 | ···················· | ···················· | ···················· | ···················· | ···················· | ···················· | |
| Q | 781 hCoV-19/Wales/PHWC-485737/2020\|EPI_ISL_628212\|2020 | ···················· | ···················· | ···················· | ···················· | ···················· | ···················· | |
| Q | 782 hCoV-19/Wales/PHWC-485746/2020\|EPI_ISL_628213\|2020 | ···················· | ···················· | ···················· | ···················· | ···················· | ···················· | |
| Q | 783 hCoV-19/Wales/PHWC-485755/2020\|EPI_ISL_628214\|2020 | ···················· | ···················· | ···················· | ···················· | ···················· | ···················· | |
| Q | 784 hCoV-19/Wales/PHWC-485764/2020\|EPI_ISL_628215\|2020 | ···················· | ···················· | ···················· | ···················· | ···················· | ···················· | |
| Q | 785 hCoV-19/Wales/PHWC-485773/2020\|EPI_ISL_628216\|2020 | ···················· | ···················· | ···················· | ···················· | ···················· | ···················· | |
| Q | 786 hCoV-19/Wales/PHWC-485791/2020\|EPI_ISL_628217\|2020 | ···················· | ···················· | ···················· | ···················· | ···················· | ···················· | |
| Q | 787 hCoV-19/Wales/PHWC-4857A0/2020\|EPI_ISL_628218\|2020 | ···················· | ···················· | ···················· | ···················· | ···················· | ···················· | |
| Q | 788 hCoV-19/Wales/PHWC-4857BF/2020\|EPI_ISL_628219\|2020 | ···················· | ···················· | ···················· | ···················· | ···················· | ···················· | |
| Q | 789 hCoV-19/Wales/PHWC-4857CE/2020\|EPI_ISL_628220\|2020 | ···················· | ···················· | ···················· | ···················· | ···················· | ···················· | |
| Q | 790 hCoV-19/Wales/PHWC-4857DD/2020\|EPI_ISL_628221\|2020 | ···················· | ···················· | ···················· | ···················· | ···················· | ···················· | |
| Q | 791 hCoV-19/Wales/PHWC-4857EC/2020\|EPI_ISL_628222\|2020 | ···················· | ···················· | ···················· | ···················· | ···················· | ···················· | |
| Q | 792 hCoV-19/Wales/PHWC-4857FB/2020\|EPI_ISL_628223\|2020 | ···················· | ···················· | ···················· | ···················· | ···················· | ···················· | |
| Q | 793 hCoV-19/Wales/PHWC-485807/2020\|EPI_ISL_628224\|2020 | ···················· | ···················· | ···················· | ···················· | ···················· | ···················· | |
| Q | 794 hCoV-19/Wales/PHWC-485816/2020\|EPI_ISL_628225\|2020 | ···················· | ···················· | ···················· | ···················· | ···················· | ···················· | |
| Q | 795 hCoV-19/Wales/PHWC-485825/2020\|EPI_ISL_628226\|2020 | ···················· | ···················· | ···················· | ···················· | ···················· | ···················· | |
| Q | 796 hCoV-19/Wales/PHWC-485843/2020\|EPI_ISL_628227\|2020 | ···················· | ···················· | ···················· | ···················· | ···················· | ···················· | |
| Q | 797 hCoV-19/Wales/PHWC-485861/2020\|EPI_ISL_628228\|2020 | ···················· | ···················· | ···················· | ···················· | ···················· | ···················· | |
| Q | 798 hCoV-19/Wales/PHWC-485870/2020\|EPI_ISL_628229\|2020 | ···················· | ···················· | ···················· | ···················· | ···················· | ···················· | |
| Q | 799 hCoV-19/Wales/PHWC-48588F/2020\|EPI_ISL_628230\|2020 | ···················· | ···················· | ···················· | ···················· | ···················· | ···················· | |
| Q | 800 hCoV-19/Wales/PHWC-4858AD/2020\|EPI_ISL_628231\|2020 | ···················· | ···················· | ···················· | ···················· | ···················· | ···················· | |
| Q | 801 hCoV-19/Wales/PHWC-4858CB/2020\|EPI_ISL_628232\|2020 | ···················· | ···················· | ···················· | ···················· | ···················· | ···················· | |
| Q | 802 hCoV-19/Wales/PHWC-4858DA/2020\|EPI_ISL_628233\|2020 | ···················· | ···················· | ···················· | ···················· | ···················· | ···················· | |
| Q | 803 hCoV-19/Wales/PHWC-4858F8/2020\|EPI_ISL_628234\|2020 | ···················· | ···················· | ···················· | ···················· | ···················· | ···················· | |
| Q | 804 hCoV-19/Wales/PHWC-485922/2020\|EPI_ISL_628235\|2020 | ···················· | ···················· | ···················· | ···················· | ···················· | ···················· | |
| Q | 805 hCoV-19/Wales/PHWC-48595F/2020\|EPI_ISL_628236\|2020 | ···················· | ···················· | ···················· | ···················· | ···················· | ···················· | |
| Q | 806 hCoV-19/Wales/PHWC-48596E/2020\|EPI_ISL_628237\|2020 | ···················· | ···················· | ···················· | ···················· | ···················· | ···················· | |
| Q | 807 hCoV-19/Wales/PHWC-48599B/2020\|EPI_ISL_628238\|2020 | ···················· | ···················· | ···················· | ···················· | ···················· | ···················· | |
| Q | 808 hCoV-19/Wales/PHWC-4859B9/2020\|EPI_ISL_628239\|2020 | ···················· | ···················· | ···················· | ···················· | ···················· | ···················· | |
| Q | 809 hCoV-19/England/MILK-AC9036/2020\|EPI_ISL_629500\|20 | ···················· | ···················· | ···················· | ···················· | ···················· | ···················· | |
| Q | 810 hCoV-19/England/MILK-ACE7A5/2020\|EPI_ISL_629501\|20 | ···················· | ···················· | ···················· | ···················· | ···················· | ···················· | |
| Q | 811 hCoV-19/England/MILK-AC819E/2020\|EPI_ISL_629502\|20 | ···················· | ···················· | ···················· | ···················· | ···················· | ···················· | |
| Q | 812 hCoV-19/England/MILK-ACE9EB/2020\|EPI_ISL_629504\|20 | ···················· | ···················· | ···················· | ···················· | ···················· | ···················· | |
| Q | 813 hCoV-19/England/MILK-ACF1E2/2020\|EPI_ISL_629505\|20 | ···················· | ···················· | ···················· | ···················· | ···················· | ···················· | |
| Q | 814 hCoV-19/England/MILK-ACEABB/2020\|EPI_ISL_629506\|20 | ···················· | ···················· | ···················· | ···················· | ···················· | ···················· | |
| Q | 815 hCoV-19/England/MILK-ACEA15/2020\|EPI_ISL_629507\|20 | ···················· | ···················· | ···················· | ···················· | ···················· | ···················· | |

| | | | | | | | | |
|---|---|---|---|---|---|---|---|---|
| Q | 816 | hCoV-19/England/MILK-ACF9DB/2020\|EPI_ISL_629508\|20 | ···················· | ···················· | ···················· | ···················· | ···················· | ···················· |
| Q | 817 | hCoV-19/England/MILK-ACE7D2/2020\|EPI_ISL_629510\|20 | ···················· | ···················· | ···················· | ···················· | ···················· | ···················· |
| Q | 818 | hCoV-19/England/MILK-AC92F4/2020\|EPI_ISL_629511\|20 | ···················· | ···················· | ···················· | ···················· | ···················· | ···················· |
| Q | 819 | hCoV-19/England/MILK-AC8F1C/2020\|EPI_ISL_629513\|20 | ···················· | ···················· | ···················· | ···················· | ···················· | ···················· |
| Q | 820 | hCoV-19/England/MILK-ACEA7F/2020\|EPI_ISL_629514\|20 | ···················· | ···················· | ···················· | ···················· | ···················· | ···················· |
| Q | 821 | hCoV-19/England/MILK-AC7FA4/2020\|EPI_ISL_629515\|20 | ···················· | ···················· | ···················· | ···················· | ···················· | ···················· |
| Q | 822 | hCoV-19/England/MILK-ACEB5E/2020\|EPI_ISL_629517\|20 | ···················· | ···················· | ···················· | ···················· | ···················· | ···················· |
| Q | 823 | hCoV-19/England/MILK-AC8170/2020\|EPI_ISL_629518\|20 | ···················· | ···················· | ···················· | ···················· | ···················· | ···················· |
| Q | 824 | hCoV-19/England/MILK-ACE5D8/2020\|EPI_ISL_629519\|20 | ···················· | ···················· | ···················· | ···················· | ···················· | ···················· |
| Q | 825 | hCoV-19/England/MILK-AC7F2C/2020\|EPI_ISL_629521\|20 | ···················· | ···················· | ···················· | ···················· | ···················· | ···················· |
| Q | 826 | hCoV-19/England/MILK-ACF425/2020\|EPI_ISL_629522\|20 | ···················· | ···················· | ···················· | ···················· | ···················· | ···················· |
| Q | 827 | hCoV-19/England/MILK-ACEE46/2020\|EPI_ISL_629523\|20 | ···················· | ···················· | ···················· | ···················· | ···················· | ···················· |
| Q | 828 | hCoV-19/England/MILK-ACEA60/2020\|EPI_ISL_629524\|20 | ···················· | ···················· | ···················· | ···················· | ···················· | ···················· |
| Q | 829 | hCoV-19/England/MILK-AC80A0/2020\|EPI_ISL_629525\|20 | ···················· | ···················· | ···················· | ···················· | ···················· | ···················· |
| Q | 830 | hCoV-19/England/MILK-ACE9DC/2020\|EPI_ISL_629527\|20 | ···················· | ···················· | ···················· | ···················· | ···················· | ···················· |
| Q | 831 | hCoV-19/England/MILK-AC8EB5/2020\|EPI_ISL_629529\|20 | ···················· | ···················· | ···················· | ···················· | ···················· | ···················· |
| Q | 832 | hCoV-19/England/MILK-ACFA50/2020\|EPI_ISL_629530\|20 | ···················· | ···················· | ···················· | ···················· | ···················· | ···················· |
| Q | 833 | hCoV-19/England/MILK-ACEFDA/2020\|EPI_ISL_629531\|20 | ···················· | ···················· | ···················· | ···················· | ···················· | ···················· |
| Q | 834 | hCoV-19/England/MILK-ACE435/2020\|EPI_ISL_629535\|20 | ···················· | ···················· | ···················· | ···················· | ···················· | ···················· |
| Q | 835 | hCoV-19/England/MILK-ACF276/2020\|EPI_ISL_629536\|20 | ···················· | ···················· | ···················· | ···················· | ···················· | ···················· |
| Q | 836 | hCoV-19/England/MILK-ACF3BF/2020\|EPI_ISL_629537\|20 | ···················· | ···················· | ···················· | ···················· | ···················· | ···················· |
| Q | 837 | hCoV-19/England/MILK-AC8E3D/2020\|EPI_ISL_629538\|20 | ···················· | ···················· | ···················· | ···················· | ···················· | ···················· |
| Q | 838 | hCoV-19/England/MILK-ACF3A0/2020\|EPI_ISL_629539\|20 | ···················· | ···················· | ···················· | ···················· | ···················· | ···················· |
| Q | 839 | hCoV-19/England/MILK-AC8ED3/2020\|EPI_ISL_629540\|20 | ···················· | ···················· | ···················· | ···················· | ···················· | ···················· |
| Q | 840 | hCoV-19/England/MILK-ACEA9D/2020\|EPI_ISL_629541\|20 | ···················· | ···················· | ···················· | ···················· | ···················· | ···················· |
| Q | 841 | hCoV-19/England/MILK-AC91BB/2020\|EPI_ISL_629542\|20 | ···················· | ···················· | ···················· | ···················· | ···················· | ···················· |
| Q | 842 | hCoV-19/England/MILK-AC8DD6/2020\|EPI_ISL_629543\|20 | ···················· | ···················· | ···················· | ···················· | ···················· | ···················· |
| Q | 843 | hCoV-19/England/MILK-ACEBB8/2020\|EPI_ISL_629545\|20 | ···················· | ···················· | ···················· | ···················· | ···················· | ···················· |
| Q | 844 | hCoV-19/England/MILK-AC95CE/2020\|EPI_ISL_629547\|20 | ···················· | ·A·················· | ···················· | ···················· | ···················· | ···A················ |
| Q | 845 | hCoV-19/England/MILK-AC9090/2020\|EPI_ISL_629550\|20 | ···················· | ···················· | ···················· | ···················· | ···················· | ···················· |
| Q | 846 | hCoV-19/England/MILK-AC89A5/2020\|EPI_ISL_629551\|20 | ···················· | ···················· | ···················· | ···················· | ···················· | ···················· |
| Q | 847 | hCoV-19/England/MILK-AC9203/2020\|EPI_ISL_629552\|20 | ···················· | ···················· | ···················· | ···················· | ···················· | ···················· |
| Q | 848 | hCoV-19/England/MILK-ACE769/2020\|EPI_ISL_629555\|20 | ···················· | ···················· | ···················· | ···················· | ···················· | ···················· |
| Q | 849 | hCoV-19/England/MILK-AC8E6A/2020\|EPI_ISL_629556\|20 | ···················· | ···················· | ···················· | ···················· | ···················· | ···················· |
| Q | 850 | hCoV-19/England/MILK-AC81F8/2020\|EPI_ISL_629558\|20 | ···················· | ···················· | ···················· | ···················· | ···················· | ···················· |
| Q | 851 | hCoV-19/England/MILK-ACEECE/2020\|EPI_ISL_629560\|20 | ···················· | ···················· | ···················· | ···················· | ···················· | ···················· |
| Q | 852 | hCoV-19/England/MILK-AC8DF4/2020\|EPI_ISL_629561\|20 | ···················· | ···················· | ···················· | ···················· | ···················· | ···················· |
| Q | 853 | hCoV-19/England/MILK-ACEEDD/2020\|EPI_ISL_629564\|20 | ···················· | ···················· | ···················· | ···················· | ····T··············· | ···················· |
| Q | 854 | hCoV-19/England/MILK-ACE6F3/2020\|EPI_ISL_629565\|20 | ···················· | ···················· | ···················· | ···················· | ···················· | ···················· |
| Q | 855 | hCoV-19/England/MILK-ACF48F/2020\|EPI_ISL_629566\|20 | ···················· | ···················· | ···················· | ···················· | ···················· | ···················· |
| Q | 856 | hCoV-19/England/MILK-ACE74B/2020\|EPI_ISL_629569\|20 | ···················· | ···················· | ···················· | ···················· | ···················· | ···················· |
| Q | 857 | hCoV-19/England/ALDP-A6745C/2020\|EPI_ISL_630500\|20 | ···················· | ···················· | ···················· | ···················· | ···················· | ···················· |
| Q | 858 | hCoV-19/England/ALDP-A66C28/2020\|EPI_ISL_630501\|20 | ···················· | ···················· | ···················· | ···················· | ···················· | ···················· |
| Q | 859 | hCoV-19/England/ALDP-A66E9B/2020\|EPI_ISL_630502\|20 | ···················· | ···················· | ···················· | ···················· | ···················· | ···················· |
| Q | 860 | hCoV-19/England/ALDP-A677BD/2020\|EPI_ISL_630503\|20 | ···················· | ···················· | ···················· | ···················· | ···················· | ···················· |
| Q | 861 | hCoV-19/England/ALDP-A66BFE/2020\|EPI_ISL_630504\|20 | ···················· | ···················· | ···················· | ···················· | ···················· | ···················· |
| Q | 862 | hCoV-19/England/ALDP-A670C1/2020\|EPI_ISL_630505\|20 | ···················· | ···················· | ···················· | ···················· | ···················· | ···················· |
| Q | 863 | hCoV-19/Wales/ALDP-A67832/2020\|EPI_ISL_630506\|2020 | ···················· | ···················· | ···················· | ···················· | ······T············· | ···················· |
| Q | 864 | hCoV-19/England/ALDP-A66E5F/2020\|EPI_ISL_630507\|20 | ···················· | ···················· | ···················· | ···················· | ···················· | ···················· |
| Q | 865 | hCoV-19/England/ALDP-A67C45/2020\|EPI_ISL_630509\|20 | ···················· | ···················· | ···················· | ···················· | ···················· | ···················· |
| Q | 866 | hCoV-19/England/ALDP-A67270/2020\|EPI_ISL_630510\|20 | ···················· | ···················· | ···················· | ···················· | ···················· | ···················· |
| Q | 867 | hCoV-19/England/ALDP-A67CBE/2020\|EPI_ISL_630511\|20 | ···················· | ···················· | ···················· | ···················· | ···················· | ···················· |
| Q | 868 | hCoV-19/England/ALDP-A672CB/2020\|EPI_ISL_630512\|20 | ···················· | ···················· | ···················· | ···················· | ···················· | ···················· |
| Q | 869 | hCoV-19/England/ALDP-A66F7A/2020\|EPI_ISL_630514\|20 | ···················· | ···················· | ···················· | ···················· | ···················· | ···················· |
| Q | 870 | hCoV-19/England/ALDP-A67C18/2020\|EPI_ISL_630515\|20 | ···················· | ·A·················· | ···················· | ···················· | ···················· | ···A················ |
| Q | 871 | hCoV-19/England/ALDP-A6689D/2020\|EPI_ISL_630516\|20 | ···················· | ···················· | ···················· | ···················· | ···················· | ···················· |
| Q | 872 | hCoV-19/England/ALDP-A67207/2020\|EPI_ISL_630517\|20 | ···················· | ···················· | ···················· | ···················· | ···················· | ···················· |
| Q | 873 | hCoV-19/England/ALDP-A66842/2020\|EPI_ISL_630518\|20 | ···················· | ·A·················· | ···················· | ···················· | ···················· | ···A················ |
| Q | 874 | hCoV-19/England/ALDP-A6794E/2020\|EPI_ISL_630519\|20 | ···················· | ···················· | ···················· | ···················· | ···················· | ···················· |
| Q | 875 | hCoV-19/England/ALDP-A6787E/2020\|EPI_ISL_630520\|20 | ···················· | ···················· | ···················· | ···················· | ···················· | ···················· |
| Q | 876 | hCoV-19/England/ALDP-A689A7/2020\|EPI_ISL_630521\|20 | ···················· | ···················· | ···················· | ···················· | ···················· | ···················· |
| Q | 877 | hCoV-19/England/ALDP-A667EB/2020\|EPI_ISL_630522\|20 | ···················· | ·A·················· | ···················· | ···················· | ···················· | ···A················ |
| Q | 878 | hCoV-19/England/ALDP-A686CE/2020\|EPI_ISL_630523\|20 | ···················· | ···················· | ···················· | ···················· | ···················· | ···················· |
| Q | 879 | hCoV-19/England/ALDP-A66860/2020\|EPI_ISL_630524\|20 | ···················· | ···················· | ···················· | ···················· | ···················· | ···················· |
| Q | 880 | hCoV-19/England/ALDP-A66FA7/2020\|EPI_ISL_630525\|20 | ···················· | ···················· | ···················· | ···················· | ···················· | ···················· |
| Q | 881 | hCoV-19/England/ALDP-A668D9/2020\|EPI_ISL_630526\|20 | ···················· | ···················· | ···················· | ···················· | ···················· | ···················· |
| Q | 882 | hCoV-19/England/ALDP-A68D9C/2020\|EPI_ISL_630528\|20 | ···················· | ···················· | ···················· | ···················· | ···················· | ···················· |
| Q | 883 | hCoV-19/England/ALDP-A6750E/2020\|EPI_ISL_630530\|20 | ···················· | ···················· | ···················· | ···················· | ···················· | ···················· |
| Q | 884 | hCoV-19/England/ALDP-A68D7E/2020\|EPI_ISL_630531\|20 | ···················· | ···················· | ···················· | ···················· | ···················· | ···················· |
| Q | 885 | hCoV-19/England/ALDP-A67AC3/2020\|EPI_ISL_630532\|20 | ···················· | ···················· | ···················· | ···················· | ···················· | ···················· |
| Q | 886 | hCoV-19/England/ALDP-A676ED/2020\|EPI_ISL_630534\|20 | ···················· | ···················· | ···················· | ···················· | ···················· | ···················· |
| Q | 887 | hCoV-19/England/ALDP-A66C55/2020\|EPI_ISL_630535\|20 | ···················· | ···················· | ···················· | ···················· | ···················· | ···················· |
| Q | 888 | hCoV-19/England/ALDP-A66DBC/2020\|EPI_ISL_630538\|20 | ···················· | ···················· | ···················· | ···················· | ···················· | ···················· |
| Q | 889 | hCoV-19/England/ALDP-A66BA3/2020\|EPI_ISL_630539\|20 | ···················· | ···················· | ···················· | ···················· | ···················· | ···················· |
| Q | 890 | hCoV-19/England/ALDP-A67692/2020\|EPI_ISL_630541\|20 | ···················· | ···················· | ···················· | ···················· | ···················· | ···················· |
| Q | 891 | hCoV-19/England/ALDP-A679D5/2020\|EPI_ISL_630542\|20 | ···················· | ···················· | ···················· | ···················· | ···················· | ···················· |
| Q | 892 | hCoV-19/England/ALDP-A66D8F/2020\|EPI_ISL_630543\|20 | ···················· | ···················· | ···················· | ···················· | ···················· | ···················· |
| Q | 893 | hCoV-19/England/ALDP-A6735F/2020\|EPI_ISL_630544\|20 | ···················· | ···················· | ···················· | ···················· | ···················· | ···················· |
| Q | 894 | hCoV-19/England/ALDP-A6737D/2020\|EPI_ISL_630548\|20 | ···················· | ···················· | ···················· | ···················· | ···················· | ···················· |
| Q | 895 | hCoV-19/England/ALDP-A67726/2020\|EPI_ISL_630549\|20 | ···················· | ···················· | ···················· | ···················· | ···················· | ···················· |

| | | | | | | | |
|---|---|---|---|---|---|---|---|
| Q | 896 hCoV-19/England/ALDP-A67595/2020\|EPI_ISL_630550\|20 | .................... | .................... | ..................... | .................... | .................... | .................... |
| Q | 897 hCoV-19/England/ALDP-A68DC9/2020\|EPI_ISL_630551\|20 | .................... | .................... | ..................... | .................... | .................... | .................... |
| Q | 898 hCoV-19/England/ALDP-A671EC/2020\|EPI_ISL_630552\|20 | .................... | .................... | ..................... | .................... | .................... | .................... |
| Q | 899 hCoV-19/England/ALDP-A66AC4/2020\|EPI_ISL_630553\|20 | .................... | .................... | ..................... | .................... | .................... | .................... |
| Q | 900 hCoV-19/England/ALDP-A6697C/2020\|EPI_ISL_630554\|20 | .................... | .A.................. | ..................... | .................... | .................... | ...A................ |
| Q | 901 hCoV-19/USA/NY-NYCPHL-000274/2020\|EPI_ISL_631800\|2 | .................... | .................... | ..................... | .................... | .................... | .................... |
| Q | 902 hCoV-19/USA/NY-NYCPHL-000275/2020\|EPI_ISL_631801\|2 | .................... | .................... | ..................... | .................... | .................... | .................... |
| Q | 903 hCoV-19/USA/NY-NYCPHL-000276/2020\|EPI_ISL_631802\|2 | .................... | .................... | ..................... | .................... | .................... | .................... |
| Q | 904 hCoV-19/USA/NY-NYCPHL-000277/2020\|EPI_ISL_631803\|2 | .................... | .................... | ..................... | .................... | .................... | .................... |
| Q | 905 hCoV-19/USA/NY-NYCPHL-000278/2020\|EPI_ISL_631804\|2 | .................... | .................... | ..................... | .................... | .................... | .................... |
| Q | 906 hCoV-19/USA/NY-NYCPHL-000279/2020\|EPI_ISL_631805\|2 | .................... | .................... | ..................... | .................... | .................... | .................... |
| Q | 907 hCoV-19/USA/NY-NYCPHL-000280/2020\|EPI_ISL_631806\|2 | .................... | .................... | ..................... | .................... | .................... | .................... |
| Q | 908 hCoV-19/USA/NY-NYCPHL-000281/2020\|EPI_ISL_631807\|2 | .................... | .................... | ..................... | .................... | .................... | .................... |
| Q | 909 hCoV-19/USA/NY-NYCPHL-000282/2020\|EPI_ISL_631808\|2 | .................... | .................... | ..................... | .................... | .................... | .................... |
| Q | 910 hCoV-19/USA/NY-NYCPHL-000283/2020\|EPI_ISL_631809\|2 | .................... | .................... | ..................... | .................... | .................... | .................... |
| Q | 911 hCoV-19/USA/NY-NYCPHL-000284/2020\|EPI_ISL_631810\|2 | .................... | .................... | ..................... | .................... | .................... | .................... |
| Q | 912 hCoV-19/USA/NY-NYCPHL-000285/2020\|EPI_ISL_631811\|2 | .................... | .................... | ..................... | .................... | .................... | .................... |
| Q | 913 hCoV-19/USA/NY-NYCPHL-000286/2020\|EPI_ISL_631812\|2 | .................... | .................... | ..................... | .................... | .................... | .................... |
| Q | 914 hCoV-19/USA/NY-NYCPHL-000287/2020\|EPI_ISL_631813\|2 | .................... | .................... | ..................... | .................... | .................... | .................... |
| Q | 915 hCoV-19/USA/NY-NYCPHL-000289/2020\|EPI_ISL_631815\|2 | .................... | .................... | ..................... | .................... | .................... | .................... |
| Q | 916 hCoV-19/USA/NY-NYCPHL-000290/2020\|EPI_ISL_631816\|2 | .................... | .................... | ..................... | .................... | .................... | .................... |
| Q | 917 hCoV-19/USA/NY-NYCPHL-000291/2020\|EPI_ISL_631817\|2 | .................... | .................... | ..................... | .................... | .................... | .................... |
| Q | 918 hCoV-19/USA/NY-NYCPHL-000292/2020\|EPI_ISL_631818\|2 | .................... | .................... | ..................... | .................... | .................... | .................... |
| Q | 919 hCoV-19/USA/NY-NYCPHL-000293/2020\|EPI_ISL_631819\|2 | .................... | .................... | ..................... | .................... | .................... | .................... |
| Q | 920 hCoV-19/USA/NY-NYCPHL-000294/2020\|EPI_ISL_631820\|2 | .................... | .................... | ..................... | .................... | .................... | .................... |
| Q | 921 hCoV-19/USA/NY-NYCPHL-000296/2020\|EPI_ISL_631822\|2 | .................... | .................... | ..................... | .................... | .................... | .................... |
| Q | 922 hCoV-19/USA/NY-NYCPHL-000298/2020\|EPI_ISL_631825\|2 | .................... | .................... | ..................... | .................... | .................... | .................... |
| Q | 923 hCoV-19/USA/NY-NYCPHL-000299/2020\|EPI_ISL_631826\|2 | .................... | .................... | ..................... | .................... | .................... | .................... |
| Q | 924 hCoV-19/USA/NY-NYCPHL-000300/2020\|EPI_ISL_631827\|2 | .................... | .................... | ..................... | .................... | .................... | .................... |
| Q | 925 hCoV-19/USA/NY-NYCPHL-000301/2020\|EPI_ISL_631828\|2 | .................... | .................... | ..................... | .................... | .................... | .................... |
| Q | 926 hCoV-19/USA/NY-NYCPHL-000302/2020\|EPI_ISL_631829\|2 | .................... | .................... | ..................... | .................... | .................... | .................... |
| Q | 927 hCoV-19/USA/NY-NYCPHL-000303/2020\|EPI_ISL_631830\|2 | .................... | .................... | ..................... | .................... | .................... | .................... |
| Q | 928 hCoV-19/USA/NY-NYCPHL-000304/2020\|EPI_ISL_631831\|2 | .................... | .................... | ..................... | .................... | .................... | .................... |
| Q | 929 hCoV-19/USA/NY-NYCPHL-000305/2020\|EPI_ISL_631832\|2 | .................... | .................... | ..................... | .................... | .................... | .................... |
| Q | 930 hCoV-19/USA/NY-NYCPHL-000306/2020\|EPI_ISL_631833\|2 | .................... | .................... | ..................... | .................... | .................... | .................... |
| Q | 931 hCoV-19/USA/NY-NYCPHL-000308/2020\|EPI_ISL_631835\|2 | .................... | .................... | ..................... | .................... | .................... | .................... |
| Q | 932 hCoV-19/USA/NY-NYCPHL-000309/2020\|EPI_ISL_631836\|2 | .................... | .................... | ..................... | .................... | .................... | .................... |
| Q | 933 hCoV-19/USA/NY-NYCPHL-000310/2020\|EPI_ISL_631837\|2 | .................... | .................... | ..................... | .................... | .................... | .................... |
| Q | 934 hCoV-19/USA/NY-NYCPHL-000311/2020\|EPI_ISL_631838\|2 | .................... | .................... | ..................... | .................... | .................... | .................... |
| Q | 935 hCoV-19/USA/NY-NYCPHL-000314/2020\|EPI_ISL_631840\|2 | .................... | .................... | ..................... | .................... | .................... | .................... |
| Q | 936 hCoV-19/USA/NY-NYCPHL-000315/2020\|EPI_ISL_631841\|2 | .................... | .................... | ..................... | .................... | .................... | .................... |
| Q | 937 hCoV-19/USA/NY-NYCPHL-000319/2020\|EPI_ISL_631843\|2 | .................... | .................... | ..................... | .................... | .................... | .................... |
| Q | 938 hCoV-19/USA/NY-NYCPHL-000332/2020\|EPI_ISL_631845\|2 | .................... | .................... | ..................... | .................... | .................... | .................... |
| Q | 939 hCoV-19/USA/NY-NYCPHL-000334/2020\|EPI_ISL_631847\|2 | .................... | .................... | ..................... | .................... | .................... | .................... |
| Q | 940 hCoV-19/USA/NY-NYCPHL-000335/2020\|EPI_ISL_631848\|2 | .................... | .................... | ..................... | .................... | .................... | .................... |
| Q | 941 hCoV-19/USA/NY-NYCPHL-000337/2020\|EPI_ISL_631849\|2 | .................... | .................... | ..................... | .................... | .................... | .................... |
| Q | 942 hCoV-19/USA/NY-NYCPHL-000339/2020\|EPI_ISL_631851\|2 | .................... | .................... | ..................... | .................... | .................... | .................... |
| Q | 943 hCoV-19/USA/NY-NYCPHL-000340/2020\|EPI_ISL_631852\|2 | .................... | .................... | ..................... | .................... | .................... | .................... |
| Q | 944 hCoV-19/USA/NY-NYCPHL-000341/2020\|EPI_ISL_631853\|2 | .................... | .................... | ..................... | .................... | .................... | .................... |
| Q | 945 hCoV-19/USA/NY-NYCPHL-000342/2020\|EPI_ISL_631854\|2 | .................... | .................... | ..................... | .................... | .................... | .................... |
| Q | 946 hCoV-19/USA/NY-NYCPHL-000343/2020\|EPI_ISL_631855\|2 | .................... | .................... | ..................... | .................... | .................... | .................... |
| Q | 947 hCoV-19/USA/NY-NYCPHL-000353/2020\|EPI_ISL_631860\|2 | .................... | .................... | ..................... | .................... | .................... | .................... |
| Q | 948 hCoV-19/USA/NY-NYCPHL-000354/2020\|EPI_ISL_631861\|2 | .................... | .................... | ..................... | .................... | .................... | .................... |
| Q | 949 hCoV-19/USA/NY-NYCPHL-000356/2020\|EPI_ISL_631862\|2 | .................... | .................... | ..................... | .................... | .................... | .................... |
| Q | 950 hCoV-19/USA/NY-NYCPHL-000357/2020\|EPI_ISL_631863\|2 | .................... | .................... | ..................... | .................... | .................... | .................... |
| Q | 951 hCoV-19/USA/NY-NYCPHL-000360/2020\|EPI_ISL_631864\|2 | .................... | .................... | ..................... | .................... | .................... | .................... |
| Q | 952 hCoV-19/USA/NY-NYCPHL-000363/2020\|EPI_ISL_631866\|2 | .................... | .................... | ..................... | .................... | .................... | .................... |
| Q | 953 hCoV-19/USA/NY-NYCPHL-000364/2020\|EPI_ISL_631867\|2 | .................... | .................... | ..................... | .................... | .................... | .................... |
| Q | 954 hCoV-19/USA/NY-NYCPHL-000379/2020\|EPI_ISL_631873\|2 | .................... | .................... | ..................... | .................... | .................... | .................... |
| Q | 955 hCoV-19/USA/NY-NYCPHL-000383/2020\|EPI_ISL_631875\|2 | .................... | .................... | ..................... | .................... | .................... | .................... |
| Q | 956 hCoV-19/USA/NY-NYCPHL-000390/2020\|EPI_ISL_631876\|2 | .................... | .................... | ..................... | .................... | .................... | .................... |
| Q | 957 hCoV-19/USA/NY-NYCPHL-000394/2020\|EPI_ISL_631879\|2 | .................... | .................... | ..................... | .................... | .................... | .................... |
| Q | 958 hCoV-19/USA/NY-NYCPHL-000396/2020\|EPI_ISL_631880\|2 | .................... | .................... | ..................... | .................... | .................... | .................... |
| Q | 959 hCoV-19/USA/NY-NYCPHL-000397/2020\|EPI_ISL_631881\|2 | .................... | .................... | ..................... | .................... | .................... | .................... |
| Q | 960 hCoV-19/USA/NY-NYCPHL-000398/2020\|EPI_ISL_631882\|2 | .................... | .................... | ..................... | .................... | .................... | .................... |
| Q | 961 hCoV-19/USA/NY-NYCPHL-000399/2020\|EPI_ISL_631883\|2 | .................... | .................... | ..................... | .................... | .................... | .................... |
| Q | 962 hCoV-19/USA/NY-NYCPHL-000401/2020\|EPI_ISL_631885\|2 | .................... | .................... | ..................... | .................... | .................... | .................... |
| Q | 963 hCoV-19/England/QEUH-AD486D/2020\|EPI_ISL_634020\|20 | .................... | .................... | ..................... | .................... | .................... | .................... |
| Q | 964 hCoV-19/England/QEUH-AD487C/2020\|EPI_ISL_634021\|20 | .................... | .................... | ..................... | .................... | .................... | .................... |
| Q | 965 hCoV-19/England/QEUH-AD494C/2020\|EPI_ISL_634022\|20 | .................... | .................... | ..................... | .................... | .................... | .................... |
| Q | 966 hCoV-19/England/QEUH-AD298A/2020\|EPI_ISL_634023\|20 | .................... | .................... | ..................... | .................... | .................... | .................... |
| Q | 967 hCoV-19/England/QEUH-AD0719/2020\|EPI_ISL_634024\|20 | .................... | .................... | ..................... | .................... | ....T............... | .................... |
| Q | 968 hCoV-19/England/QEUH-AD243E/2020\|EPI_ISL_634025\|20 | .................... | .................... | ..................... | .................... | .................... | .................... |
| Q | 969 hCoV-19/England/QEUH-AD07EC/2020\|EPI_ISL_634026\|20 | .................... | .................... | ..................... | .................... | .................... | .................... |
| Q | 970 hCoV-19/England/QEUH-AD2076/2020\|EPI_ISL_634027\|20 | .................... | .................... | ..................... | .................... | .................... | .................... |
| Q | 971 hCoV-19/England/QEUH-AD04E5/2020\|EPI_ISL_634028\|20 | .................... | .................... | ..................... | .................... | .................... | .................... |
| Q | 972 hCoV-19/England/QEUH-AD0CFC/2020\|EPI_ISL_634029\|20 | .................... | .................... | ..................... | .................... | .................... | .................... |
| Q | 973 hCoV-19/England/QEUH-AD4821/2020\|EPI_ISL_634030\|20 | .................... | .................... | ..................... | .................... | .................... | .................... |
| Q | 974 hCoV-19/England/QEUH-AD3C44/2020\|EPI_ISL_634034\|20 | .................... | .................... | ..................... | .................... | .................... | .................... |
| Q | 975 hCoV-19/England/QEUH-AD2C90/2020\|EPI_ISL_634035\|20 | .................... | .................... | ..................... | .................... | .................... | .................... |

| | | | | | | | | |
|---|---|---|---|---|---|---|---|---|
| Q | 976 | hCoV-19/England/QEUH-AD2B57/2020\|EPI_ISL_634036\|20 | ···················· | ···················· | ···················· | ···················· | ···················· | ···················· |
| Q | 977 | hCoV-19/England/QEUH-AD053D/2020\|EPI_ISL_634037\|20 | ···················· | ···················· | ···················· | ···················· | ···················· | ···················· |
| Q | 978 | hCoV-19/England/QEUH-AD5064/2020\|EPI_ISL_634038\|20 | ···················· | ···················· | ···················· | ···················· | ···················· | ···················· |
| Q | 979 | hCoV-19/England/QEUH-AD3743/2020\|EPI_ISL_634039\|20 | ···················· | ···················· | ···········T········ | ···················· | ···················· | ···················· |
| Q | 980 | hCoV-19/England/QEUH-AD229E/2020\|EPI_ISL_634080\|20 | ···················· | ···················· | ···················· | ···················· | ···················· | ···················· |
| Q | 981 | hCoV-19/England/QEUH-AD50A0/2020\|EPI_ISL_634081\|20 | ···················· | ···················· | ···················· | ···················· | ···················· | ···················· |
| Q | 982 | hCoV-19/England/QEUH-AD40DE/2020\|EPI_ISL_634082\|20 | ···················· | ···················· | ···················· | ···················· | ···················· | ···················· |
| Q | 983 | hCoV-19/England/QEUH-AD3734/2020\|EPI_ISL_634083\|20 | ···················· | ···················· | ···················· | ···················· | ···················· | ···················· |
| Q | 984 | hCoV-19/England/QEUH-AD4108/2020\|EPI_ISL_634084\|20 | ···················· | ···················· | ···················· | ···················· | ···················· | ···················· |
| Q | 985 | hCoV-19/England/QEUH-AD0755/2020\|EPI_ISL_634085\|20 | ···················· | ···················· | ···················· | ···················· | ···················· | ···················· |
| Q | 986 | hCoV-19/England/QEUH-AD3804/2020\|EPI_ISL_634086\|20 | ···················· | ···················· | ···················· | ···················· | ···················· | ···················· |
| Q | 987 | hCoV-19/England/QEUH-AD05E2/2020\|EPI_ISL_634087\|20 | ···················· | ···················· | ···················· | ···················· | ···················· | ···················· |
| Q | 988 | hCoV-19/England/QEUH-AD34A6/2020\|EPI_ISL_634088\|20 | ···················· | ···················· | ···················· | ···················· | ···················· | ···················· |
| Q | 989 | hCoV-19/England/QEUH-AD3C62/2020\|EPI_ISL_634089\|20 | ···················· | ···················· | ···················· | ···················· | ···················· | ···················· |
| Q | 990 | hCoV-19/England/QEUH-AD2B84/2020\|EPI_ISL_634090\|20 | ···················· | ···················· | ···················· | ···················· | ···················· | ···················· |
| Q | 991 | hCoV-19/England/QEUH-AD48F4/2020\|EPI_ISL_634091\|20 | ···················· | ···················· | ···················· | ···················· | ···················· | ···················· |
| Q | 992 | hCoV-19/England/QEUH-AD03AC/2020\|EPI_ISL_634093\|20 | ···················· | ···················· | ···················· | ···················· | ···················· | ···················· |
| Q | 993 | hCoV-19/England/QEUH-AD519E/2020\|EPI_ISL_634095\|20 | ···················· | ···················· | ···················· | ···················· | ···················· | ···················· |
| Q | 994 | hCoV-19/England/QEUH-AD4636/2020\|EPI_ISL_634096\|20 | ···················· | ···················· | ···················· | ···················· | ···················· | ···················· |
| Q | 995 | hCoV-19/England/QEUH-AD51AD/2020\|EPI_ISL_634097\|20 | ···················· | ···················· | ···················· | ···················· | ···················· | ···················· |
| Q | 996 | hCoV-19/England/QEUH-AD5204/2020\|EPI_ISL_634099\|20 | ···················· | ···················· | ···················· | ···················· | ···················· | ···················· |
| Q | 997 | hCoV-19/USA/CA-ALSR-2109/2020\|EPI_ISL_635350\|2020- | ···················· | ···················· | ···················· | ···················· | ···················· | ···················· |
| Q | 998 | hCoV-19/USA/CA-ALSR-2114/2020\|EPI_ISL_635352\|2020- | ···················· | ·A·················· | ···················· | ···················· | ···················· | ···················· |
| Q | 999 | hCoV-19/USA/CA-ALSR-2115/2020\|EPI_ISL_635353\|2020- | ···················· | ·A·················· | ···················· | ···················· | ···················· | ···················· |
| Q | 1000 | hCoV-19/USA/CA-ALSR-2116/2020\|EPI_ISL_635354\|2020- | ···················· | ·A·················· | ···················· | ···················· | ···················· | ···················· |
| Q | 1001 | hCoV-19/USA/CA-ALSR-2118/2020\|EPI_ISL_635355\|2020- | ···················· | ···················· | ···················· | ···················· | ···················· | ···················· |
| Q | 1002 | hCoV-19/USA/CA-ALSR-2121/2020\|EPI_ISL_635356\|2020- | ···················· | ·A·················· | ···················· | ···················· | ···················· | ···················· |
| Q | 1003 | hCoV-19/USA/CA-ALSR-2127/2020\|EPI_ISL_635357\|2020- | ···················· | ···················· | ···················· | ···················· | ···················· | ···················· |
| Q | 1004 | hCoV-19/USA/CA-ALSR-2128/2020\|EPI_ISL_635358\|2020- | ···················· | ·A·················· | ···················· | ···················· | ···················· | ···················· |
| Q | 1005 | hCoV-19/USA/CA-ALSR-2129/2020\|EPI_ISL_635359\|2020- | ···················· | ···················· | ···················· | ···················· | ···················· | ···················· |
| Q | 1006 | hCoV-19/USA/CA-ALSR-2132/2020\|EPI_ISL_635360\|2020- | ···················· | ·A·················· | ···················· | ···················· | ···················· | ···················· |
| Q | 1007 | hCoV-19/USA/CA-ALSR-2163/2020\|EPI_ISL_635366\|2020- | ···················· | ·A·················· | ···················· | ···················· | ···················· | ···················· |
| Q | 1008 | hCoV-19/USA/CA-ALSR-2168/2020\|EPI_ISL_635369\|2020- | ···················· | ·A·················· | ···················· | ···················· | ···················· | ···················· |
| Q | 1014 | hCoV-19/England/C200021007/2020\|EPI_ISL_636686\|202 | ···················· | ···················· | ···················· | ···················· | ···················· | ···················· |
| Q | 1015 | hCoV-19/England/C200021209/2020\|EPI_ISL_636687\|202 | ···················· | ···················· | ···················· | ···················· | ···················· | ···················· |
| Q | 1016 | hCoV-19/England/C200024017/2020\|EPI_ISL_636688\|202 | ···················· | ···················· | ···················· | ···················· | ···················· | ···················· |
| Q | 1017 | hCoV-19/England/C200024019/2020\|EPI_ISL_636689\|202 | ···················· | ···················· | ···················· | ···················· | ···················· | ···················· |
| Q | 1018 | hCoV-19/England/C200028985/2020\|EPI_ISL_636690\|202 | ···················· | ···················· | ···················· | ···················· | ···················· | ···················· |
| Q | 1019 | hCoV-19/England/C200028987/2020\|EPI_ISL_636691\|202 | ···················· | ···················· | ···················· | ···················· | ···················· | ···················· |
| Q | 1020 | hCoV-19/England/C200028988/2020\|EPI_ISL_636692\|202 | ···················· | ···················· | ···················· | ···················· | ···················· | ···················· |
| Q | 1021 | hCoV-19/England/C200028989/2020\|EPI_ISL_636693\|202 | ···················· | ···················· | ···················· | ···················· | ···················· | ···················· |
| Q | 1022 | hCoV-19/England/C200028991/2020\|EPI_ISL_636694\|202 | ···················· | ···················· | ···················· | ···················· | ···················· | ···················· |
| Q | 1023 | hCoV-19/England/C200028992/2020\|EPI_ISL_636695\|202 | ···················· | ···················· | ···················· | ···················· | ···················· | ···················· |
| Q | 1024 | hCoV-19/England/C200029066/2020\|EPI_ISL_636699\|202 | ···················· | ···················· | ···················· | ···················· | ···················· | ···················· |
| Q | 1025 | hCoV-19/Wales/PHWC-484E60/2020\|EPI_ISL_628120\|2020 | ···················· | ···················· | ···················· | ···················· | ···················· | ···················· |
| Q | 1026 | hCoV-19/Wales/PHWC-484E7F/2020\|EPI_ISL_628121\|2020 | ···················· | ···················· | ···················· | ···················· | ···················· | ···················· |
| Q | 1027 | hCoV-19/Wales/PHWC-484E8E/2020\|EPI_ISL_628122\|2020 | ···················· | ···················· | ···················· | ···················· | ···················· | ···················· |
| Q | 1029 | hCoV-19/Wales/PHWC-484EAC/2020\|EPI_ISL_628124\|2020 | ···················· | ···················· | ···················· | ···················· | ···················· | ···················· |
| Q | 1030 | hCoV-19/Wales/PHWC-484EBB/2020\|EPI_ISL_628125\|2020 | ···················· | ···················· | ···················· | ···················· | ···················· | ···················· |
| Q | 1031 | hCoV-19/Wales/PHWC-484ED9/2020\|EPI_ISL_628126\|2020 | ···················· | ···················· | ···················· | ···················· | ···················· | ···················· |
| Q | 1032 | hCoV-19/Wales/PHWC-484F4F/2020\|EPI_ISL_628127\|2020 | ···················· | ···················· | ···················· | ···················· | ···················· | ···················· |
| Q | 1033 | hCoV-19/Wales/PHWC-484F6D/2020\|EPI_ISL_628128\|2020 | ···················· | ···················· | ···················· | ···················· | ···················· | ···················· |
| Q | 1034 | hCoV-19/Wales/PHWC-484F7C/2020\|EPI_ISL_628129\|2020 | ···················· | ···················· | ···················· | ···················· | ···················· | ···················· |
| Q | 1035 | hCoV-19/Wales/PHWC-484F8B/2020\|EPI_ISL_628130\|2020 | ···················· | ···················· | ···················· | ···················· | ···················· | ···················· |
| Q | 1036 | hCoV-19/Wales/PHWC-484F9A/2020\|EPI_ISL_628131\|2020 | ···················· | ···················· | ···················· | ···················· | ···················· | ···················· |
| Q | 1037 | hCoV-19/Wales/PHWC-484FA9/2020\|EPI_ISL_628132\|2020 | ···················· | ···················· | ···················· | ···················· | ···················· | ···················· |
| Q | 1038 | hCoV-19/Wales/PHWC-484FC7/2020\|EPI_ISL_628133\|2020 | ···················· | ···················· | ···················· | ···················· | ···················· | ···················· |
| Q | 1040 | hCoV-19/Wales/PHWC-484FF4/2020\|EPI_ISL_628135\|2020 | ···················· | ···················· | ···················· | ···················· | ···················· | ···················· |
| Q | 1041 | hCoV-19/Wales/PHWC-48500F/2020\|EPI_ISL_628136\|2020 | ···················· | ···················· | ···················· | ···················· | ···················· | ···················· |
| Q | 1042 | hCoV-19/Wales/PHWC-48502D/2020\|EPI_ISL_628137\|2020 | ···················· | ···················· | ···················· | ···················· | ···················· | ···················· |
| Q | 1043 | hCoV-19/Wales/PHWC-48504B/2020\|EPI_ISL_628138\|2020 | ···················· | ···················· | ···················· | ···················· | ···················· | ···················· |
| Q | 1044 | hCoV-19/Wales/PHWC-485078/2020\|EPI_ISL_628139\|2020 | ···················· | ···················· | ···················· | ···················· | ···················· | ···················· |
| Q | 1045 | hCoV-19/Wales/PHWC-485087/2020\|EPI_ISL_628140\|2020 | ···················· | ···················· | ···················· | ···················· | ···················· | ···················· |
| Q | 1046 | hCoV-19/Wales/PHWC-485096/2020\|EPI_ISL_628141\|2020 | ···················· | ···················· | ···················· | ···················· | ···················· | ···················· |
| Q | 1047 | hCoV-19/Wales/PHWC-4850A5/2020\|EPI_ISL_628142\|2020 | ···················· | ···················· | ···················· | ···················· | ···················· | ···················· |
| Q | 1048 | hCoV-19/Wales/PHWC-4850B4/2020\|EPI_ISL_628143\|2020 | ···················· | ···················· | ···················· | ···················· | ···················· | ···················· |
| Q | 1049 | hCoV-19/Wales/PHWC-4850C3/2020\|EPI_ISL_628144\|2020 | ···················· | ···················· | ···················· | ···················· | ···················· | ···················· |
| Q | 1050 | hCoV-19/Wales/PHWC-4850D2/2020\|EPI_ISL_628145\|2020 | ···················· | ···················· | ···················· | ···················· | ···················· | ···················· |
| Q | 1051 | hCoV-19/Wales/PHWC-4850E1/2020\|EPI_ISL_628146\|2020 | ···················· | ···················· | ···················· | ···················· | ···················· | ···················· |
| Q | 1052 | hCoV-19/Wales/PHWC-4850F0/2020\|EPI_ISL_628147\|2020 | ···················· | ···················· | ···················· | ···················· | ···················· | ···················· |
| Q | 1053 | hCoV-19/Wales/PHWC-48510C/2020\|EPI_ISL_628148\|2020 | ···················· | ···················· | ···················· | ···················· | ···················· | ···················· |
| Q | 1054 | hCoV-19/Wales/PHWC-48512A/2020\|EPI_ISL_628149\|2020 | ···················· | ···················· | ···················· | ···················· | ···················· | ···················· |
| Q | 1055 | hCoV-19/Wales/PHWC-485139/2020\|EPI_ISL_628150\|2020 | ···················· | ···················· | ···················· | ···················· | ···················· | ···················· |
| Q | 1056 | hCoV-19/Wales/PHWC-485148/2020\|EPI_ISL_628151\|2020 | ···················· | ···················· | ···················· | ···················· | ···················· | ···················· |
| Q | 1057 | hCoV-19/Wales/PHWC-485166/2020\|EPI_ISL_628152\|2020 | ···················· | ···················· | ···················· | ···················· | ···················· | ···················· |
| Q | 1058 | hCoV-19/Wales/PHWC-485175/2020\|EPI_ISL_628153\|2020 | ···················· | ···················· | ···················· | ···················· | ···················· | ···················· |
| Q | 1059 | hCoV-19/Wales/PHWC-485193/2020\|EPI_ISL_628154\|2020 | ···················· | ···················· | ···················· | ···················· | ···················· | ···················· |
| Q | 1060 | hCoV-19/Wales/PHWC-4851A2/2020\|EPI_ISL_628155\|2020 | ···················· | ···················· | ···················· | ···················· | ···················· | ···················· |
| Q | 1061 | hCoV-19/Wales/PHWC-4851B1/2020\|EPI_ISL_628156\|2020 | ···················· | ···················· | ···················· | ···················· | ···················· | ···················· |
| Q | 1062 | hCoV-19/Wales/PHWC-4851FD/2020\|EPI_ISL_628157\|2020 | ···················· | ···················· | ···················· | ···················· | ···················· | ···················· |

| | | | | | | | |
|---|---|---|---|---|---|---|---|
| Q | 1063 hCoV-19/Wales/PHWC-485218/2020\|EPI_ISL_628158\|2020 | ···················· | ···················· | ····················· | ···················· | ···················· | ···················· |
| Q | 1064 hCoV-19/Wales/PHWC-485227/2020\|EPI_ISL_628159\|2020 | ···················· | ···················· | ····················· | ···················· | ···················· | ···················· |
| Q | 1065 hCoV-19/Wales/PHWC-485245/2020\|EPI_ISL_628160\|2020 | ···················· | ···················· | ····················· | ···················· | ···················· | ···················· |
| Q | 1066 hCoV-19/Wales/PHWC-485254/2020\|EPI_ISL_628161\|2020 | ···················· | ···················· | ····················· | ···················· | ···················· | ···················· |
| Q | 1067 hCoV-19/Wales/PHWC-485263/2020\|EPI_ISL_628162\|2020 | ···················· | ···················· | ····················· | ···················· | ···················· | ···················· |
| Q | 1069 hCoV-19/Wales/PHWC-4852AF/2020\|EPI_ISL_628164\|2020 | ···················· | ···················· | ····················· | ···················· | ···················· | ···················· |
| Q | 1070 hCoV-19/Wales/PHWC-4852DC/2020\|EPI_ISL_628165\|2020 | ···················· | ···················· | ····················· | ···················· | ···················· | ···················· |
| Q | 1071 hCoV-19/Wales/PHWC-4852FA/2020\|EPI_ISL_628166\|2020 | ···················· | ···················· | ····················· | ···················· | ···················· | ···················· |
| Q | 1072 hCoV-19/Wales/PHWC-485306/2020\|EPI_ISL_628167\|2020 | ···················· | ···················· | ····················· | ···················· | ···················· | ···················· |
| Q | 1073 hCoV-19/Wales/PHWC-485315/2020\|EPI_ISL_628168\|2020 | ···················· | ···················· | ····················· | ···················· | ···················· | ···················· |
| Q | 1074 hCoV-19/Wales/PHWC-485324/2020\|EPI_ISL_628169\|2020 | ···················· | ···················· | ····················· | ···················· | ···················· | ···················· |
| Q | 1075 hCoV-19/Wales/PHWC-485342/2020\|EPI_ISL_628170\|2020 | ···················· | ···················· | ····················· | ···················· | ···················· | ···················· |
| Q | 1076 hCoV-19/Wales/PHWC-485351/2020\|EPI_ISL_628171\|2020 | ···················· | ···················· | ····················· | ···················· | ···················· | ···················· |
| Q | 1077 hCoV-19/Wales/PHWC-485360/2020\|EPI_ISL_628172\|2020 | ···················· | ···················· | ····················· | ···················· | ···················· | ···················· |
| Q | 1078 hCoV-19/Wales/PHWC-48537F/2020\|EPI_ISL_628173\|2020 | ···················· | ···················· | ····················· | ···················· | ···················· | ···················· |
| Q | 1079 hCoV-19/Wales/PHWC-48538E/2020\|EPI_ISL_628174\|2020 | ···················· | ···················· | ····················· | ···················· | ···················· | ···················· |
| Q | 1080 hCoV-19/Wales/PHWC-48539D/2020\|EPI_ISL_628175\|2020 | ···················· | ···················· | ····················· | ···················· | ···················· | ···················· |
| Q | 1081 hCoV-19/Wales/PHWC-4853AC/2020\|EPI_ISL_628176\|2020 | ···················· | ···················· | ····················· | ···················· | ···················· | ···················· |
| Q | 1082 hCoV-19/Wales/PHWC-4853BB/2020\|EPI_ISL_628177\|2020 | ···················· | ···················· | ····················· | ···················· | ···················· | ···················· |
| Q | 1083 hCoV-19/Wales/PHWC-4853CA/2020\|EPI_ISL_628178\|2020 | ···················· | ···················· | ····················· | ···················· | ···················· | ···················· |
| Q | 1084 hCoV-19/Wales/PHWC-4853E8/2020\|EPI_ISL_628179\|2020 | ···················· | ···················· | ····················· | ···················· | ···················· | ···················· |
| Q | 1085 hCoV-19/Wales/PHWC-4853F7/2020\|EPI_ISL_628180\|2020 | ···················· | ···················· | ····················· | ···················· | ···················· | ···················· |
| Q | 1086 hCoV-19/Wales/PHWC-485403/2020\|EPI_ISL_628181\|2020 | ···················· | ···················· | ····················· | ···················· | ···················· | ···················· |
| Q | 1087 hCoV-19/Wales/PHWC-485412/2020\|EPI_ISL_628182\|2020 | ···················· | ···················· | ····················· | ···················· | ···················· | ···················· |
| Q | 1089 hCoV-19/Wales/PHWC-48546D/2020\|EPI_ISL_628184\|2020 | ···················· | ···················· | ····················· | ···················· | ···················· | ···················· |
| Q | 1090 hCoV-19/Wales/PHWC-4854B8/2020\|EPI_ISL_628185\|2020 | ···················· | ···················· | ····················· | ···················· | ···················· | ···················· |
| Q | 1091 hCoV-19/Wales/PHWC-4854F4/2020\|EPI_ISL_628186\|2020 | ···················· | ···················· | ····················· | ···················· | ···················· | ···················· |
| Q | 1092 hCoV-19/Wales/PHWC-48551F/2020\|EPI_ISL_628187\|2020 | ···················· | ···················· | ····················· | ···················· | ···················· | ···················· |
| Q | 1093 hCoV-19/Wales/PHWC-48552E/2020\|EPI_ISL_628188\|2020 | ···················· | ···················· | ····················· | ···················· | ···················· | ···················· |
| Q | 1094 hCoV-19/Wales/PHWC-48554C/2020\|EPI_ISL_628189\|2020 | ···················· | ···················· | ····················· | ···················· | ···················· | ···················· |
| Q | 1095 hCoV-19/Wales/PHWC-48555B/2020\|EPI_ISL_628190\|2020 | ···················· | ···················· | ····················· | ···················· | ···················· | ···················· |
| Q | 1096 hCoV-19/Wales/PHWC-48556A/2020\|EPI_ISL_628191\|2020 | ···················· | ···················· | ····················· | ···················· | ···················· | ···················· |
| Q | 1097 hCoV-19/Wales/PHWC-485579/2020\|EPI_ISL_628192\|2020 | ···················· | ···················· | ····················· | ···················· | ···················· | ···················· |
| Q | 1098 hCoV-19/Wales/PHWC-485588/2020\|EPI_ISL_628193\|2020 | ···················· | ···················· | ····················· | ···················· | ···················· | ···················· |
| Q | 1099 hCoV-19/Wales/PHWC-485597/2020\|EPI_ISL_628194\|2020 | ···················· | ···················· | ····················· | ···················· | ···················· | ···················· |
| Q | 1100 hCoV-19/Wales/PHWC-4855A6/2020\|EPI_ISL_628195\|2020 | ···················· | ···················· | ····················· | ···················· | ···················· | ···················· |
| Q | 1101 hCoV-19/Wales/PHWC-4855C4/2020\|EPI_ISL_628196\|2020 | ···················· | ···················· | ····················· | ···················· | ···················· | ···················· |
| Q | 1102 hCoV-19/Wales/PHWC-4855E2/2020\|EPI_ISL_628197\|2020 | ···················· | ···················· | ····················· | ···················· | ···················· | ···················· |
| Q | 1103 hCoV-19/Wales/PHWC-4855F1/2020\|EPI_ISL_628198\|2020 | ···················· | ···················· | ····················· | ···················· | ···················· | ···················· |
| Q | 1104 hCoV-19/Wales/PHWC-48560D/2020\|EPI_ISL_628199\|2020 | ···················· | ···················· | ····················· | ···················· | ···················· | ···················· |
| Q | 1105 hCoV-19/England/MILK-AC8240/2020\|EPI_ISL_629451\|20 | ···················· | ···················· | ····················· | ···················· | ···················· | ···················· |
| Q | 1106 hCoV-19/England/MILK-ACE7E1/2020\|EPI_ISL_629452\|20 | ···················· | ···················· | ····················· | ···················· | ···················· | ···················· |
| Q | 1107 hCoV-19/England/MILK-ACE2C2/2020\|EPI_ISL_629453\|20 | ···················· | ···················· | ····················· | ···················· | ···················· | ···················· |
| Q | 1108 hCoV-19/England/MILK-ACF504/2020\|EPI_ISL_629455\|20 | ···················· | ···················· | ····················· | ···················· | ···················· | ···················· |
| Q | 1109 hCoV-19/Wales/MILK-AC84E0/2020\|EPI_ISL_629456\|2020 | ···················· | ···················· | ····················· | ···················· | ···················· | ···················· |
| Q | 1110 hCoV-19/England/MILK-AC89B4/2020\|EPI_ISL_629457\|20 | ···················· | ···················· | ····················· | ···················· | ···················· | ···················· |
| Q | 1111 hCoV-19/England/MILK-AC8565/2020\|EPI_ISL_629458\|20 | ···················· | ···················· | ····················· | ···················· | ···················· | ···················· |
| Q | 1112 hCoV-19/England/MILK-ACE6B7/2020\|EPI_ISL_629460\|20 | ···················· | ···················· | ····················· | ···················· | ···················· | ···················· |
| Q | 1113 hCoV-19/England/MILK-ACF7C2/2020\|EPI_ISL_629461\|20 | ···················· | ···················· | ····················· | ···················· | ···················· | ···················· |
| Q | 1114 hCoV-19/England/MILK-AC89C3/2020\|EPI_ISL_629463\|20 | ···················· | ···················· | ····················· | ···················· | ···················· | ···················· |
| Q | 1115 hCoV-19/England/MILK-ACF2C1/2020\|EPI_ISL_629464\|20 | ···················· | ·A·················· | ····················· | ···················· | ···················· | ··A················· |
| Q | 1116 hCoV-19/England/MILK-ACE778/2020\|EPI_ISL_629465\|20 | ···················· | ···················· | ····················· | ···················· | ·····T·············· | ···················· |
| Q | 1117 hCoV-19/England/MILK-ACEA06/2020\|EPI_ISL_629467\|20 | ···················· | ···················· | ····················· | ···················· | ···················· | ···················· |
| Q | 1118 hCoV-19/England/MILK-ACE875/2020\|EPI_ISL_629468\|20 | ···················· | ···················· | ····················· | ···················· | ···················· | ···················· |
| Q | 1119 hCoV-19/England/MILK-AC89F0/2020\|EPI_ISL_629469\|20 | ···················· | ···················· | ····················· | ···················· | ···················· | ···················· |
| Q | 1120 hCoV-19/England/MILK-ACE7C3/2020\|EPI_ISL_629470\|20 | ···················· | ···················· | ····················· | ···················· | ···················· | ···················· |
| Q | 1121 hCoV-19/England/MILK-ACEFBC/2020\|EPI_ISL_629471\|20 | ···················· | ···················· | ····················· | ···················· | ···················· | ···················· |
| Q | 1122 hCoV-19/England/MILK-AC9133/2020\|EPI_ISL_629472\|20 | ···················· | ···················· | ····················· | ···················· | ···················· | ···················· |
| Q | 1123 hCoV-19/England/MILK-AC8028/2020\|EPI_ISL_629473\|20 | ···················· | ···················· | ····················· | ···················· | ···················· | ···················· |
| Q | 1124 hCoV-19/England/MILK-ACF434/2020\|EPI_ISL_629474\|20 | ···················· | ···················· | ····················· | ···················· | ···················· | ···················· |
| Q | 1125 hCoV-19/England/MILK-AC7FE0/2020\|EPI_ISL_629475\|20 | ···················· | ···················· | ····················· | ···················· | ···················· | ···················· |
| Q | 1126 hCoV-19/England/MILK-AC8FFE/2020\|EPI_ISL_629477\|20 | ···················· | ···················· | ····················· | ···················· | ···················· | ···················· |
| Q | 1127 hCoV-19/England/MILK-ACF13D/2020\|EPI_ISL_629480\|20 | ···················· | ·A·················· | ····················· | ···················· | ···················· | ··A················· |
| Q | 1128 hCoV-19/England/MILK-ACFA23/2020\|EPI_ISL_629481\|20 | ···················· | ···················· | ····················· | ···················· | ···················· | ···················· |
| Q | 1129 hCoV-19/England/MILK-ACE59C/2020\|EPI_ISL_629482\|20 | ···················· | ···················· | ····················· | ···················· | ···················· | ···················· |
| Q | 1130 hCoV-19/England/MILK-AC7FFF/2020\|EPI_ISL_629483\|20 | ···················· | ···················· | ····················· | ···················· | ···················· | ···················· |
| Q | 1131 hCoV-19/England/MILK-ACF3EC/2020\|EPI_ISL_629485\|20 | ···················· | ···················· | ····················· | ···················· | ···················· | ···················· |
| Q | 1132 hCoV-19/England/MILK-AC81E9/2020\|EPI_ISL_629486\|20 | ···················· | ···················· | ····················· | ···················· | ···················· | ···················· |
| Q | 1133 hCoV-19/England/MILK-ACEDFE/2020\|EPI_ISL_629487\|20 | ···················· | ···················· | ····················· | ···················· | ···················· | ···················· |
| Q | 1134 hCoV-19/England/MILK-AC91AC/2020\|EPI_ISL_629488\|20 | ···················· | ···················· | ····················· | ···················· | ···················· | ···················· |
| Q | 1135 hCoV-19/England/MILK-AC7FB3/2020\|EPI_ISL_629489\|20 | ···················· | ···················· | ····················· | ···················· | ···················· | ···················· |
| Q | 1136 hCoV-19/England/MILK-ACE9FA/2020\|EPI_ISL_629491\|20 | ···················· | ···················· | ····················· | ···················· | ···T················ | ···················· |
| Q | 1137 hCoV-19/England/MILK-AC8EE2/2020\|EPI_ISL_629492\|20 | ···················· | ···················· | ····················· | ···················· | ···················· | ···················· |
| Q | 1138 hCoV-19/England/MILK-AC8A0C/2020\|EPI_ISL_629493\|20 | ···················· | ···················· | ····················· | ···················· | ···················· | ···················· |
| Q | 1139 hCoV-19/England/MILK-AC8B90/2020\|EPI_ISL_629494\|20 | ···················· | ···················· | ····················· | ···················· | ···················· | ···················· |
| Q | 1140 hCoV-19/England/MILK-AC8082/2020\|EPI_ISL_629496\|20 | ···················· | ···················· | ····················· | ···················· | ···················· | ···················· |
| Q | 1141 hCoV-19/England/MILK-AC8E1F/2020\|EPI_ISL_629497\|20 | ···················· | ···················· | ····················· | ···················· | ·····T·············· | ···················· |
| Q | 1142 hCoV-19/England/MILK-AC8222/2020\|EPI_ISL_629498\|20 | ···················· | ···················· | ····················· | ···················· | ···················· | ···················· |
| Q | 1143 hCoV-19/England/ALDP-A676B0/2020\|EPI_ISL_630434\|20 | ···················· | ···················· | ····················· | ···················· | ···················· | ···················· |
| Q | 1144 hCoV-19/England/ALDP-A68761/2020\|EPI_ISL_630436\|20 | ···················· | ·A·················· | ····················· | ···················· | ···················· | ···················· |

Q 1145 hCoV-19/England/ALDP-A669C7/2020|EPI_ISL_630437|20 ···················· ···················· ····················· ···················· ···················· ····················
Q 1146 hCoV-19/England/ALDP-A67340/2020|EPI_ISL_630438|20 ···················· ·A·················· ····················· ···················· ···················· ····················
Q 1147 hCoV-19/England/ALDP-A66A5B/2020|EPI_ISL_630439|20 ···················· ···················· ····················· ···················· ···················· ····················
Q 1148 hCoV-19/England/ALDP-A6688E/2020|EPI_ISL_630440|20 ···················· ·A·················· ····················· ···················· ···················· ····················
Q 1149 hCoV-19/England/ALDP-A6752C/2020|EPI_ISL_630441|20 ···················· ···················· ····················· ···················· ···················· ····················
Q 1150 hCoV-19/England/ALDP-A66A88/2020|EPI_ISL_630442|20 ···················· ···················· ····················· ···················· ···················· ····················
Q 1151 hCoV-19/England/ALDP-A67225/2020|EPI_ISL_630444|20 ···················· ···················· ····················· ···················· ···················· ····················
Q 1152 hCoV-19/England/ALDP-A66851/2020|EPI_ISL_630445|20 ···················· ···················· ····················· ···················· ···················· ····················
Q 1153 hCoV-19/England/ALDP-A6747A/2020|EPI_ISL_630446|20 ···················· ···················· ····················· ···················· ···················· ····················
Q 1154 hCoV-19/England/ALDP-A671DD/2020|EPI_ISL_630447|20 ···················· ···················· ····················· ···················· ···················· ····················
Q 1155 hCoV-19/England/ALDP-A66B0D/2020|EPI_ISL_630448|20 ···················· ···················· ····················· ···················· ···················· ····················
Q 1156 hCoV-19/England/ALDP-A68DF6/2020|EPI_ISL_630449|20 ···················· ···················· ····················· ···················· ···················· ····················
Q 1157 hCoV-19/England/ALDP-A66DF8/2020|EPI_ISL_630450|20 ···················· ···················· ····················· ···················· ···················· ····················
Q 1158 hCoV-19/England/ALDP-A67401/2020|EPI_ISL_630451|20 ···················· ···················· ····················· ···················· ···················· ····················
Q 1159 hCoV-19/England/ALDP-A67A3C/2020|EPI_ISL_630452|20 ···················· ···················· ····················· ···················· ···················· ····················
Q 1160 hCoV-19/England/ALDP-A66833/2020|EPI_ISL_630453|20 ···················· ···················· ····················· ···················· ···················· ····················
Q 1161 hCoV-19/England/ALDP-A66A4C/2020|EPI_ISL_630454|20 ···················· ···················· ····················· ···················· ···················· ····················
Q 1162 hCoV-19/England/ALDP-A66CBF/2020|EPI_ISL_630456|20 ···················· ···················· ····················· ···················· ···················· ····················
Q 1163 hCoV-19/England/ALDP-A66E8C/2020|EPI_ISL_630457|20 ···················· ···················· ····················· ···················· ···················· ····················
Q 1164 hCoV-19/England/ALDP-A66B49/2020|EPI_ISL_630458|20 ···················· ···················· ····················· ···················· ···················· ····················
Q 1165 hCoV-19/England/ALDP-A66AE2/2020|EPI_ISL_630459|20 ···················· ···················· ····················· ···················· ···················· ····················
Q 1166 hCoV-19/England/ALDP-A67498/2020|EPI_ISL_630460|20 ···················· ···················· ····················· ···················· ···················· ····················
Q 1167 hCoV-19/England/ALDP-A670A3/2020|EPI_ISL_630461|20 ···················· ···················· ····················· ···················· ···················· ····················
Q 1168 hCoV-19/England/ALDP-A6696D/2020|EPI_ISL_630462|20 ···················· ·A·················· ····················· ···················· ···················· ···A················
Q 1169 hCoV-19/England/ALDP-A6695E/2020|EPI_ISL_630463|20 ···················· ···················· ····················· ···················· ···················· ····················
Q 1170 hCoV-19/England/ALDP-A679C6/2020|EPI_ISL_630464|20 ···················· ···················· ····················· ···················· ···················· ····················
Q 1171 hCoV-19/England/ALDP-A67489/2020|EPI_ISL_630465|20 ···················· ···················· ····················· ···················· ···················· ····················
Q 1172 hCoV-19/England/ALDP-A67261/2020|EPI_ISL_630466|20 ···················· ···················· ····················· ···················· ···················· ····················
Q 1173 hCoV-19/England/ALDP-A66DAD/2020|EPI_ISL_630467|20 ···················· ···················· ····················· ···················· ···················· ····················
Q 1174 hCoV-19/England/ALDP-A673AA/2020|EPI_ISL_630468|20 ···················· ···················· ····················· ···················· ···················· ····················
Q 1175 hCoV-19/England/ALDP-A6896B/2020|EPI_ISL_630469|20 ···················· ···················· ····················· ···················· ······T············· ····················
Q 1176 hCoV-19/England/ALDP-A6736E/2020|EPI_ISL_630470|20 ···················· ···················· ····················· ···················· ···················· ····················
Q 1177 hCoV-19/England/ALDP-A66B58/2020|EPI_ISL_630471|20 ···················· ···················· ····················· ···················· ···················· ····················
Q 1178 hCoV-19/England/ALDP-A670D0/2020|EPI_ISL_630472|20 ···················· ···················· ····················· ···················· ···················· ····················
Q 1179 hCoV-19/England/ALDP-A67C81/2020|EPI_ISL_630473|20 ···················· ···················· ····················· ···················· ···················· ····················
Q 1180 hCoV-19/England/ALDP-A66DE9/2020|EPI_ISL_630475|20 ···················· ···················· ····················· ···················· ···················· ····················
Q 1181 hCoV-19/England/ALDP-A68752/2020|EPI_ISL_630476|20 ···················· ···················· ····················· ···················· ···················· ····················
Q 1182 hCoV-19/England/ALDP-A66DCB/2020|EPI_ISL_630477|20 ···················· ···················· ····················· ···················· ···················· ····················
Q 1183 hCoV-19/England/ALDP-A67A4B/2020|EPI_ISL_630478|20 ···················· ···················· ····················· ···················· ···················· ····················
Q 1184 hCoV-19/England/ALDP-A676FC/2020|EPI_ISL_630479|20 ···················· ···················· ····················· ···················· ···················· ····················
Q 1185 hCoV-19/England/ALDP-A67CEB/2020|EPI_ISL_630480|20 ···················· ·A·················· ····················· ···················· ···················· ···A················
Q 1186 hCoV-19/England/ALDP-A6703A/2020|EPI_ISL_630481|20 ···················· ···················· ····················· ···················· ···················· ····················
Q 1187 hCoV-19/England/ALDP-A67137/2020|EPI_ISL_630482|20 ···················· ···················· ····················· ···················· ···················· ····················
Q 1188 hCoV-19/England/ALDP-A66F89/2020|EPI_ISL_630484|20 ···················· ···················· ····················· ···················· ···················· ····················
Q 1189 hCoV-19/England/ALDP-A66C37/2020|EPI_ISL_630485|20 ···················· ···················· ····················· ···················· ···················· ····················
Q 1190 hCoV-19/England/ALDP-A67771/2020|EPI_ISL_630486|20 ···················· ···················· ····················· ···················· ···················· ····················
Q 1191 hCoV-19/England/ALDP-A68DE7/2020|EPI_ISL_630487|20 ···················· ···················· ····················· ···················· ···················· ····················
Q 1192 hCoV-19/England/ALDP-A66B76/2020|EPI_ISL_630488|20 ···················· ···················· ····················· ···················· ···················· ····················
Q 1193 hCoV-19/England/ALDP-A66B1C/2020|EPI_ISL_630490|20 ···················· ···················· ····················· ···················· ···················· ····················
Q 1194 hCoV-19/England/ALDP-A679B7/2020|EPI_ISL_630491|20 ···················· ···················· ····················· ···················· ···················· ····················
Q 1195 hCoV-19/England/ALDP-A68682/2020|EPI_ISL_630492|20 ···················· ···················· ····················· ···················· ···················· ····················
Q 1196 hCoV-19/England/ALDP-A67AA5/2020|EPI_ISL_630493|20 ···················· ···················· ····················· ···················· ···················· ····················
Q 1197 hCoV-19/England/ALDP-A66DDA/2020|EPI_ISL_630494|20 ···················· ···················· ····················· ···················· ···················· ····················
Q 1198 hCoV-19/England/ALDP-A68707/2020|EPI_ISL_630495|20 ···················· ···················· ····················· ···················· ···················· ····················
Q 1199 hCoV-19/England/ALDP-A669F4/2020|EPI_ISL_630496|20 ···················· ···················· ····················· ···················· ···················· ····················
Q 1200 hCoV-19/England/ALDP-A677AE/2020|EPI_ISL_630497|20 ···················· ···················· ····················· ···················· ···················· ····················
Q 1201 hCoV-19/England/ALDP-A66E22/2020|EPI_ISL_630498|20 ···················· ···················· ····················· ···················· ···················· ····················
Q 1202 hCoV-19/England/ALDP-A67067/2020|EPI_ISL_630499|20 ···················· ···················· ····················· ···················· ···················· ····················
Q 1203 hCoV-19/USA/NY-NYCPHL-000237/2020|EPI_ISL_631766|2 ···················· ···················· ····················· ···················· ···················· ····················
Q 1204 hCoV-19/USA/NY-NYCPHL-000238/2020|EPI_ISL_631767|2 ···················· ···················· ····················· ···················· ···················· ····················
Q 1205 hCoV-19/USA/NY-NYCPHL-000241/2020|EPI_ISL_631768|2 ···················· ···················· ····················· ···················· ···················· ····················
Q 1206 hCoV-19/USA/NY-NYCPHL-000242/2020|EPI_ISL_631769|2 ···················· ···················· ····················· ···················· ···················· ····················
Q 1207 hCoV-19/USA/NY-NYCPHL-000243/2020|EPI_ISL_631770|2 ···················· ···················· ····················· ···················· ···················· ····················
Q 1208 hCoV-19/USA/NY-NYCPHL-000244/2020|EPI_ISL_631771|2 ···················· ···················· ····················· ···················· ···················· ····················
Q 1209 hCoV-19/USA/NY-NYCPHL-000246/2020|EPI_ISL_631772|2 ···················· ···················· ····················· ···················· ···················· ····················
Q 1210 hCoV-19/USA/NY-NYCPHL-000247/2020|EPI_ISL_631773|2 ···················· ···················· ····················· ···················· ···················· ····················
Q 1211 hCoV-19/USA/NY-NYCPHL-000248/2020|EPI_ISL_631774|2 ···················· ···················· ····················· ···················· ···················· ····················
Q 1212 hCoV-19/USA/NY-NYCPHL-000249/2020|EPI_ISL_631775|2 ···················· ···················· ····················· ···················· ···················· ····················
Q 1213 hCoV-19/USA/NY-NYCPHL-000250/2020|EPI_ISL_631776|2 ···················· ···················· ····················· ···················· ···················· ····················
Q 1214 hCoV-19/USA/NY-NYCPHL-000251/2020|EPI_ISL_631777|2 ···················· ···················· ····················· ···················· ···················· ····················
Q 1215 hCoV-19/USA/NY-NYCPHL-000252/2020|EPI_ISL_631778|2 ···················· ···················· ····················· ···················· ···················· ····················
Q 1216 hCoV-19/USA/NY-NYCPHL-000253/2020|EPI_ISL_631779|2 ···················· ···················· ····················· ···················· ···················· ····················
Q 1217 hCoV-19/USA/NY-NYCPHL-000254/2020|EPI_ISL_631780|2 ···················· ···················· ····················· ···················· ···················· ····················
Q 1218 hCoV-19/USA/NY-NYCPHL-000255/2020|EPI_ISL_631781|2 ···················· ···················· ····················· ···················· ···················· ····················
Q 1219 hCoV-19/USA/NY-NYCPHL-000256/2020|EPI_ISL_631782|2 ···················· ···················· ····················· ···················· ···················· ····················
Q 1220 hCoV-19/USA/NY-NYCPHL-000258/2020|EPI_ISL_631784|2 ···················· ···················· ····················· ···················· ···················· ····················
Q 1221 hCoV-19/USA/NY-NYCPHL-000259/2020|EPI_ISL_631785|2 ···················· ···················· ····················· ···················· ···················· ····················
Q 1222 hCoV-19/USA/NY-NYCPHL-000260/2020|EPI_ISL_631786|2 ···················· ···················· ····················· ···················· ···················· ····················
Q 1223 hCoV-19/USA/NY-NYCPHL-000261/2020|EPI_ISL_631787|2 ···················· ···················· ····················· ···················· ···················· ····················
Q 1224 hCoV-19/USA/NY-NYCPHL-000262/2020|EPI_ISL_631788|2 ···················· ···················· ····················· ···················· ···················· ····················

| | | | | | | | |
|---|---|---|---|---|---|---|---|
| Q | 1225 hCoV-19/USA/NY-NYCPHL-000263/2020\|EPI_ISL_631789\|2 | ···················· | ···················· | ····················· | ···················· | ···················· | ···················· |
| Q | 1226 hCoV-19/USA/NY-NYCPHL-000266/2020\|EPI_ISL_631792\|2 | ···················· | ···················· | ····················· | ···················· | ···················· | ···················· |
| Q | 1227 hCoV-19/USA/NY-NYCPHL-000267/2020\|EPI_ISL_631793\|2 | ···················· | ···················· | ····················· | ···················· | ···················· | ···················· |
| Q | 1228 hCoV-19/USA/NY-NYCPHL-000268/2020\|EPI_ISL_631794\|2 | ···················· | ···················· | ····················· | ···················· | ···················· | ···················· |
| Q | 1229 hCoV-19/USA/NY-NYCPHL-000270/2020\|EPI_ISL_631796\|2 | ···················· | ···················· | ····················· | ···················· | ···················· | ···················· |
| Q | 1230 hCoV-19/USA/NY-NYCPHL-000271/2020\|EPI_ISL_631797\|2 | ···················· | ···················· | ····················· | ···················· | ···················· | ···················· |
| Q | 1231 hCoV-19/USA/CA-ALSR-1996/2020\|EPI_ISL_635300\|2020- | ···················· | ·A·················· | ····················· | ···················· | ···················· | ···················· |
| Q | 1232 hCoV-19/USA/CA-ALSR-2004/2020\|EPI_ISL_635301\|2020- | ···················· | ·A·················· | ····················· | ···················· | ···················· | ···················· |
| Q | 1233 hCoV-19/USA/CA-ALSR-2005/2020\|EPI_ISL_635302\|2020- | ···················· | ···················· | ····················· | ···················· | ···················· | ···················· |
| Q | 1234 hCoV-19/USA/CA-ALSR-2010/2020\|EPI_ISL_635303\|2020- | ···················· | ·A·················· | ····················· | ···················· | ···················· | ···················· |
| Q | 1235 hCoV-19/USA/CA-ALSR-2011/2020\|EPI_ISL_635304\|2020- | ···················· | ·A·················· | ····················· | ···················· | ···················· | ···················· |
| Q | 1236 hCoV-19/USA/CA-ALSR-2012/2020\|EPI_ISL_635305\|2020- | ···················· | ·A·················· | ····················· | ···················· | ···················· | ···················· |
| Q | 1237 hCoV-19/USA/CA-ALSR-2015/2020\|EPI_ISL_635306\|2020- | ···················· | ·A·················· | ····················· | ···················· | ···················· | ···················· |
| Q | 1238 hCoV-19/USA/CA-ALSR-2051/2020\|EPI_ISL_635318\|2020- | ···················· | ·A·················· | ····················· | ···················· | ···················· | ···················· |
| Q | 1239 hCoV-19/USA/CA-ALSR-2052/2020\|EPI_ISL_635319\|2020- | ···················· | ·A·················· | ····················· | ···················· | ···················· | ···················· |
| Q | 1240 hCoV-19/USA/CA-ALSR-2053/2020\|EPI_ISL_635320\|2020- | ···················· | ·A·················· | ····················· | ···················· | ···················· | ···················· |
| Q | 1241 hCoV-19/USA/CA-ALSR-2055/2020\|EPI_ISL_635321\|2020- | ···················· | ·A·················· | ····················· | ···················· | ···················· | ···················· |
| Q | 1242 hCoV-19/USA/CA-ALSR-2056/2020\|EPI_ISL_635322\|2020- | ···················· | ·A·················· | ····················· | ···················· | ···················· | ···················· |
| Q | 1243 hCoV-19/USA/CA-ALSR-2057/2020\|EPI_ISL_635323\|2020- | ···················· | ·A·················· | ····················· | ···················· | ···················· | ···················· |
| Q | 1244 hCoV-19/USA/CA-ALSR-2058/2020\|EPI_ISL_635324\|2020- | ···················· | ·A·················· | ····················· | ···················· | ···················· | ···················· |
| Q | 1245 hCoV-19/USA/CA-ALSR-2060/2020\|EPI_ISL_635325\|2020- | ···················· | ·A·················· | ····················· | ···················· | ···················· | ···················· |
| Q | 1246 hCoV-19/USA/CA-ALSR-2061/2020\|EPI_ISL_635326\|2020- | ···················· | ·A·················· | ····················· | ···················· | ···················· | ···················· |
| Q | 1247 hCoV-19/USA/CA-ALSR-2062/2020\|EPI_ISL_635327\|2020- | ···················· | ·A·················· | ····················· | ···················· | ···················· | ···················· |
| Q | 1248 hCoV-19/USA/CA-ALSR-2063/2020\|EPI_ISL_635328\|2020- | ···················· | ·A·················· | ····················· | ···················· | ···················· | ···················· |
| Q | 1254 hCoV-19/Belgium/ULG-10644/2020\|EPI_ISL_636622\|2020 | ···················· | ···················· | ····················· | ···················· | ···················· | ···················· |
| Q | 1261 hCoV-19/Belgium/ULG-10651/2020\|EPI_ISL_636629\|2020 | ···················· | ···················· | ····················· | ···················· | ···················· | ···················· |
| Q | 1263 hCoV-19/Belgium/ULG-10654/2020\|EPI_ISL_636631\|2020 | ···················· | ···················· | ····················· | ···················· | ···················· | ···················· |
| Q | 1264 hCoV-19/Belgium/ULG-10655/2020\|EPI_ISL_636632\|2020 | ···················· | ···················· | ····················· | ···················· | ···················· | ···················· |
| Q | 1265 hCoV-19/Belgium/ULG-10656/2020\|EPI_ISL_636633\|2020 | ···················· | ···················· | ····················· | ···················· | ···················· | ···················· |
| Q | 1266 hCoV-19/Belgium/ULG-10657/2020\|EPI_ISL_636634\|2020 | ···················· | ···················· | ····················· | ···················· | ···················· | ···················· |
| Q | 1269 hCoV-19/Belgium/ULG-10660/2020\|EPI_ISL_636637\|2020 | ···················· | ···················· | ····················· | ···················· | ···················· | ···················· |
| Q | 1277 hCoV-19/Belgium/ULG-10684/2020\|EPI_ISL_636656\|2020 | ···················· | ···················· | ····················· | ···················· | ···················· | ···················· |
| Q | 1279 hCoV-19/Belgium/ULG-10686/2020\|EPI_ISL_636658\|2020 | ···················· | ···················· | ····················· | ···················· | ···················· | ···················· |
| Q | 1281 hCoV-19/Wales/PHWC-484DAF/2020\|EPI_ISL_628110\|2020 | ···················· | ···················· | ····················· | ···················· | ···················· | ···················· |
| Q | 1282 hCoV-19/Wales/PHWC-484DBE/2020\|EPI_ISL_628111\|2020 | ···················· | ···················· | ····················· | ···················· | ···················· | ···················· |
| Q | 1283 hCoV-19/Wales/PHWC-484E15/2020\|EPI_ISL_628116\|2020 | ···················· | ···················· | ····················· | ···················· | ···················· | ···················· |
| Q | 1284 hCoV-19/Wales/PHWC-484E24/2020\|EPI_ISL_628117\|2020 | ···················· | ···················· | ····················· | ···················· | ···················· | ···················· |
| Q | 1285 hCoV-19/Wales/PHWC-484E42/2020\|EPI_ISL_628118\|2020 | ···················· | ···················· | ····················· | ···················· | ···················· | ···················· |
| Q | 1286 hCoV-19/Wales/PHWC-484E51/2020\|EPI_ISL_628119\|2020 | ···················· | ···················· | ····················· | ···················· | ···················· | ···················· |
| Q | 1287 hCoV-19/England/MILK-AC7D22/2020\|EPI_ISL_629410\|20 | ···················· | ···················· | ····················· | ···················· | ···················· | ···················· |
| Q | 1288 hCoV-19/England/MILK-ACCA80/2020\|EPI_ISL_629411\|20 | ···················· | ···················· | ····················· | ···················· | ···················· | ···················· |
| Q | 1289 hCoV-19/England/MILK-ACC73E/2020\|EPI_ISL_629412\|20 | ···················· | ···················· | ····················· | ···················· | ···················· | ···················· |
| Q | 1290 hCoV-19/England/MILK-ACC0D9/2020\|EPI_ISL_629413\|20 | ···················· | ···················· | ····················· | ···················· | ···················· | ···················· |
| Q | 1291 hCoV-19/England/MILK-ACE8A2/2020\|EPI_ISL_629414\|20 | ···················· | ···················· | ····················· | ···················· | ···················· | ···················· |
| Q | 1292 hCoV-19/England/MILK-ACC358/2020\|EPI_ISL_629415\|20 | ···················· | ···················· | ····················· | ···················· | ···················· | ···················· |
| Q | 1293 hCoV-19/England/MILK-AC929A/2020\|EPI_ISL_629416\|20 | ···················· | ···················· | ····················· | ···················· | ···················· | ···················· |
| Q | 1294 hCoV-19/England/MILK-AC92E5/2020\|EPI_ISL_629417\|20 | ···················· | ···················· | ····················· | ···················· | ·····T·············· | ···················· |
| Q | 1295 hCoV-19/England/MILK-ACC75C/2020\|EPI_ISL_629418\|20 | ···················· | ···················· | ····················· | ···················· | ·····T·············· | ···················· |
| Q | 1296 hCoV-19/England/MILK-ACCA9F/2020\|EPI_ISL_629419\|20 | ···················· | ·A·················· | ····················· | ···················· | ···················· | ··A················· |
| Q | 1297 hCoV-19/England/MILK-ACC14F/2020\|EPI_ISL_629420\|20 | ···················· | ···················· | ····················· | ···················· | ·····T·············· | ···················· |
| Q | 1298 hCoV-19/England/MILK-ACBF73/2020\|EPI_ISL_629421\|20 | ···················· | ···················· | ····················· | ···················· | ···················· | ···················· |
| Q | 1299 hCoV-19/England/MILK-ACC32B/2020\|EPI_ISL_629422\|20 | ···················· | ···················· | ····················· | ···················· | ···················· | ···················· |
| Q | 1300 hCoV-19/England/MILK-ACC9ED/2020\|EPI_ISL_629423\|20 | ···················· | ···················· | ····················· | ···················· | ···················· | ···················· |
| Q | 1301 hCoV-19/England/MILK-ACC65F/2020\|EPI_ISL_629424\|20 | ···················· | ···················· | ····················· | ···················· | ···················· | ···················· |
| Q | 1302 hCoV-19/England/MILK-ACBDC4/2020\|EPI_ISL_629425\|20 | ···················· | ···················· | ····················· | ···················· | ···················· | ···················· |
| Q | 1303 hCoV-19/England/MILK-AC8617/2020\|EPI_ISL_629426\|20 | ···················· | ···················· | ····················· | ···················· | ···················· | ···················· |
| Q | 1304 hCoV-19/England/MILK-AC7CE9/2020\|EPI_ISL_629427\|20 | ···················· | ···················· | ····················· | ···················· | ···················· | ···················· |
| Q | 1305 hCoV-19/England/MILK-AC83F2/2020\|EPI_ISL_629429\|20 | ···················· | ···················· | ····················· | ···················· | ···················· | ···················· |
| Q | 1306 hCoV-19/England/MILK-ACEFCB/2020\|EPI_ISL_629441\|20 | ···················· | ···················· | ····················· | ···················· | ···················· | ···················· |
| Q | 1307 hCoV-19/England/MILK-ACF7A4/2020\|EPI_ISL_629442\|20 | ···················· | ···················· | ····················· | ···················· | ·····T·············· | ···············A···· |
| Q | 1308 hCoV-19/England/MILK-AC9027/2020\|EPI_ISL_629447\|20 | ···················· | ···················· | ····················· | ···················· | ···················· | ···················· |
| Q | 1309 hCoV-19/England/MILK-ACEB4F/2020\|EPI_ISL_629448\|20 | ···················· | ·········A·········· | ····················· | ···················· | ···················· | ···················· |
| Q | 1310 hCoV-19/England/MILK-ACF2A3/2020\|EPI_ISL_629449\|20 | ···················· | ···················· | ····················· | ···················· | ···················· | ···················· |
| Q | 1311 hCoV-19/England/ALDP-A6746B/2020\|EPI_ISL_630400\|20 | ···················· | ···················· | ····················· | ···················· | ···················· | ···················· |
| Q | 1312 hCoV-19/England/ALDP-A668BB/2020\|EPI_ISL_630401\|20 | ···················· | ···················· | ····················· | ···················· | ···················· | ···················· |
| Q | 1313 hCoV-19/England/ALDP-A68743/2020\|EPI_ISL_630402\|20 | ···················· | ···················· | ····················· | ···················· | ···················· | ···················· |
| Q | 1314 hCoV-19/England/ALDP-A67164/2020\|EPI_ISL_630403\|20 | ···················· | ···················· | ····················· | ···················· | ···················· | ···················· |
| Q | 1315 hCoV-19/England/ALDP-A66F10/2020\|EPI_ISL_630404\|20 | ···················· | ···················· | ····················· | ···················· | ···················· | ···················· |
| Q | 1316 hCoV-19/England/ALDP-A66D70/2020\|EPI_ISL_630405\|20 | ···················· | ···················· | ····················· | ···················· | ···················· | ···················· |
| Q | 1317 hCoV-19/England/ALDP-A68D23/2020\|EPI_ISL_630406\|20 | ···················· | ···················· | ····················· | ···················· | ···················· | ···················· |
| Q | 1318 hCoV-19/England/ALDP-A66CA0/2020\|EPI_ISL_630407\|20 | ···················· | ···················· | ····················· | ···················· | ···················· | ···················· |
| Q | 1319 hCoV-19/England/ALDP-A6742F/2020\|EPI_ISL_630408\|20 | ···················· | ···················· | ····················· | ···················· | ···················· | ···················· |
| Q | 1320 hCoV-19/England/ALDP-A68655/2020\|EPI_ISL_630409\|20 | ···················· | ···················· | ····················· | ···················· | ···················· | ···················· |
| Q | 1321 hCoV-19/England/ALDP-A67AB4/2020\|EPI_ISL_630410\|20 | ···················· | ···················· | ····················· | ···················· | ···················· | ···················· |
| Q | 1322 hCoV-19/England/ALDP-A679A8/2020\|EPI_ISL_630411\|20 | ···················· | ···················· | ····················· | ···················· | ···················· | ···················· |
| Q | 1323 hCoV-19/England/ALDP-A6743E/2020\|EPI_ISL_630423\|20 | ···················· | ···················· | ····················· | ···················· | ···················· | ···················· |
| Q | 1324 hCoV-19/England/ALDP-A686A0/2020\|EPI_ISL_630424\|20 | ···················· | ·A·················· | ····················· | ···················· | ···················· | ···················· |
| Q | 1325 hCoV-19/England/ALDP-A689F2/2020\|EPI_ISL_630425\|20 | ···················· | ···················· | ····················· | ···················· | ···················· | ···················· |
| Q | 1326 hCoV-19/England/ALDP-A667DC/2020\|EPI_ISL_630426\|20 | ···················· | ···················· | ····················· | ···················· | ···················· | ···················· |
| Q | 1327 hCoV-19/England/ALDP-A66C19/2020\|EPI_ISL_630427\|20 | ···················· | ···················· | ····················· | ···················· | ···················· | ···················· |

| | | | | | | | |
|---|---|---|---|---|---|---|---|
| Q | 1328 hCoV-19/England/ALDP-A66D07/2020\|EPI_ISL_630428\|20 | ···················· | ···················· | ···················· | ···················· | ···················· | ···················· |
| Q | 1329 hCoV-19/England/ALDP-A669B8/2020\|EPI_ISL_630429\|20 | ···················· | ···················· | ···················· | ···················· | ···················· | ···················· |
| Q | 1330 hCoV-19/England/ALDP-A66D34/2020\|EPI_ISL_630430\|20 | ···················· | ···················· | ···················· | ···················· | ···················· | ···················· |
| Q | 1331 hCoV-19/England/ALDP-A669A9/2020\|EPI_ISL_630431\|20 | ···················· | ·A·················· | ···················· | ···················· | ···················· | ··A················· |
| Q | 1332 hCoV-19/England/ALDP-A674B6/2020\|EPI_ISL_630432\|20 | ···················· | ···················· | ···················· | ···················· | ···················· | ···················· |
| Q | 1333 hCoV-19/England/ALDP-A66930/2020\|EPI_ISL_630433\|20 | ···················· | ···················· | ···················· | ···················· | ···················· | ···················· |
| Q | 1334 hCoV-19/USA/NY-NYCPHL-000186/2020\|EPI_ISL_631721\|2 | ···················· | ···················· | ···················· | ···················· | ···················· | ···················· |
| Q | 1335 hCoV-19/USA/NY-NYCPHL-000187/2020\|EPI_ISL_631722\|2 | ···················· | ···················· | ···················· | ···················· | ···················· | ···················· |
| Q | 1336 hCoV-19/USA/NY-NYCPHL-000189/2020\|EPI_ISL_631724\|2 | ···················· | ···················· | ···················· | ···················· | ···················· | ···················· |
| Q | 1337 hCoV-19/USA/NY-NYCPHL-000190/2020\|EPI_ISL_631725\|2 | ···················· | ···················· | ···················· | ···················· | ···················· | ···················· |
| Q | 1338 hCoV-19/USA/NY-NYCPHL-000191/2020\|EPI_ISL_631726\|2 | ···················· | ···················· | ···················· | ···················· | ···················· | ···················· |
| Q | 1339 hCoV-19/USA/NY-NYCPHL-000193/2020\|EPI_ISL_631728\|2 | ···················· | ···················· | ···················· | ···················· | ···················· | ···················· |
| Q | 1340 hCoV-19/USA/NY-NYCPHL-000195/2020\|EPI_ISL_631730\|2 | ···················· | ···················· | ···················· | ···················· | ···················· | ···················· |
| Q | 1341 hCoV-19/USA/NY-NYCPHL-000196/2020\|EPI_ISL_631731\|2 | ···················· | ···················· | ···················· | ···················· | ···················· | ···················· |
| Q | 1342 hCoV-19/USA/NY-NYCPHL-000197/2020\|EPI_ISL_631732\|2 | ···················· | ···················· | ···················· | ···················· | ···················· | ···················· |
| Q | 1343 hCoV-19/USA/NY-NYCPHL-000198/2020\|EPI_ISL_631733\|2 | ···················· | ···················· | ···················· | ···················· | ···················· | ···················· |
| Q | 1344 hCoV-19/USA/NY-NYCPHL-000201/2020\|EPI_ISL_631735\|2 | ···················· | ···················· | ···················· | ···················· | ···················· | ···················· |
| Q | 1345 hCoV-19/USA/NY-NYCPHL-000202/2020\|EPI_ISL_631736\|2 | ···················· | ···················· | ···················· | ···················· | ···················· | ···················· |
| Q | 1346 hCoV-19/USA/NY-NYCPHL-000203/2020\|EPI_ISL_631737\|2 | ···················· | ···················· | ···················· | ···················· | ···················· | ···················· |
| Q | 1347 hCoV-19/USA/NY-NYCPHL-000205/2020\|EPI_ISL_631738\|2 | ···················· | ···················· | ···················· | ···················· | ···················· | ···················· |
| Q | 1348 hCoV-19/USA/NY-NYCPHL-000206/2020\|EPI_ISL_631739\|2 | ···················· | ···················· | ···················· | ···················· | ···················· | ···················· |
| Q | 1349 hCoV-19/USA/NY-NYCPHL-000207/2020\|EPI_ISL_631740\|2 | ···················· | ···················· | ···················· | ···················· | ···················· | ···················· |
| Q | 1350 hCoV-19/USA/NY-NYCPHL-000208/2020\|EPI_ISL_631741\|2 | ···················· | ···················· | ···················· | ···················· | ···················· | ···················· |
| Q | 1351 hCoV-19/USA/NY-NYCPHL-000209/2020\|EPI_ISL_631742\|2 | ···················· | ···················· | ···················· | ···················· | ···················· | ···················· |
| Q | 1352 hCoV-19/USA/NY-NYCPHL-000224/2020\|EPI_ISL_631754\|2 | ···················· | ···················· | ···················· | ···················· | ···················· | ···················· |
| Q | 1353 hCoV-19/USA/NY-NYCPHL-000226/2020\|EPI_ISL_631755\|2 | ···················· | ···················· | ···················· | ···················· | ···················· | ···················· |
| Q | 1354 hCoV-19/USA/NY-NYCPHL-000227/2020\|EPI_ISL_631756\|2 | ···················· | ··············C····· | ···················· | ···················· | ···················· | ···················· |
| Q | 1355 hCoV-19/USA/NY-NYCPHL-000228/2020\|EPI_ISL_631757\|2 | ···················· | ···················· | ···················· | ···················· | ···················· | ···················· |
| Q | 1356 hCoV-19/USA/NY-NYCPHL-000229/2020\|EPI_ISL_631758\|2 | ···················· | ···················· | ···················· | ···················· | ···················· | ···················· |
| Q | 1357 hCoV-19/USA/NY-NYCPHL-000230/2020\|EPI_ISL_631759\|2 | ···················· | ···················· | ···················· | ···················· | ···················· | ···················· |
| Q | 1358 hCoV-19/USA/NY-NYCPHL-000231/2020\|EPI_ISL_631760\|2 | ···················· | ···················· | ···················· | ···················· | ···················· | ···················· |
| Q | 1359 hCoV-19/USA/NY-NYCPHL-000232/2020\|EPI_ISL_631761\|2 | ···················· | ···················· | ···················· | ···················· | ···················· | ···················· |
| Q | 1360 hCoV-19/USA/NY-NYCPHL-000233/2020\|EPI_ISL_631762\|2 | ···················· | ···················· | ···················· | ···················· | ···················· | ···················· |
| Q | 1361 hCoV-19/USA/NY-NYCPHL-000234/2020\|EPI_ISL_631763\|2 | ···················· | ···················· | ···················· | ···················· | ···················· | ···················· |
| Q | 1362 hCoV-19/USA/NY-NYCPHL-000235/2020\|EPI_ISL_631764\|2 | ···················· | ···················· | ···················· | ···················· | ···················· | ···················· |
| Q | 1363 hCoV-19/Slovenia/4815/2020\|EPI_ISL_635208\|2020-03- | ···················· | ···················· | ···················· | ···················· | ···················· | ···················· |
| Q | 1364 hCoV-19/Slovenia/4915/2020\|EPI_ISL_635209\|2020-03- | ···················· | ···················· | ···················· | ···················· | ···················· | ···················· |
| Q | 1365 hCoV-19/Slovenia/4996/2020\|EPI_ISL_635210\|2020-03- | ···················· | ···················· | ···················· | ···················· | ···················· | ···················· |
| Q | 1366 hCoV-19/Slovenia/5027/2020\|EPI_ISL_635211\|2020-03- | ···················· | ···················· | ···················· | ···················· | ···················· | ···················· |
| Q | 1367 hCoV-19/Slovenia/5028/2020\|EPI_ISL_635212\|2020-03- | ···················· | ···················· | ···················· | ···················· | ···················· | ···················· |
| Q | 1368 hCoV-19/Slovenia/5070/2020\|EPI_ISL_635213\|2020-03- | ···················· | ···················· | ···················· | ···················· | ···················· | ···················· |
| Q | 1369 hCoV-19/Slovenia/5091/2020\|EPI_ISL_635214\|2020-03- | ···················· | ···················· | ···················· | ···················· | ···················· | ···················· |
| Q | 1370 hCoV-19/Slovenia/5166/2020\|EPI_ISL_635215\|2020-03- | ···················· | ···················· | ···················· | ···················· | ···················· | ···················· |
| Q | 1371 hCoV-19/Slovenia/5177/2020\|EPI_ISL_635216\|2020-03- | ···················· | ···················· | ···················· | ···················· | ···················· | ···················· |
| Q | 1373 hCoV-19/Slovenia/5359/2020\|EPI_ISL_635218\|2020-03- | ···················· | ···················· | ···················· | ···················· | ···················· | ···················· |
| Q | 1374 hCoV-19/Slovenia/5551/2020\|EPI_ISL_635219\|2020-03- | ···················· | ···················· | ···················· | ···················· | ···················· | ···················· |
| Q | 1375 hCoV-19/Slovenia/5677/2020\|EPI_ISL_635220\|2020-03- | ···················· | ···················· | ···················· | ···················· | ···················· | ···················· |
| Q | 1376 hCoV-19/Slovenia/5703/2020\|EPI_ISL_635221\|2020-03- | ···················· | ···················· | ···················· | ···················· | ···················· | ···················· |
| Q | 1377 hCoV-19/Slovenia/5715/2020\|EPI_ISL_635222\|2020-03- | ···················· | ···················· | ···················· | ···················· | ···················· | ···················· |
| Q | 1378 hCoV-19/Slovenia/5716/2020\|EPI_ISL_635223\|2020-03- | ···················· | ···················· | ···················· | ···················· | ···················· | ···················· |
| Q | 1379 hCoV-19/Slovenia/5775/2020\|EPI_ISL_635224\|2020-03- | ···················· | ···················· | ···················· | ···················· | ···················· | ···················· |
| Q | 1380 hCoV-19/Slovenia/5982/2020\|EPI_ISL_635225\|2020-03- | ···················· | ···················· | ···················· | ···················· | ···················· | ···················· |
| Q | 1381 hCoV-19/Slovenia/6051/2020\|EPI_ISL_635226\|2020-03- | ···················· | ···················· | ···················· | ···················· | ···················· | ···················· |
| Q | 1382 hCoV-19/Slovenia/6098/2020\|EPI_ISL_635227\|2020-03- | ···················· | ···················· | ···················· | ···················· | ···················· | ···················· |
| Q | 1383 hCoV-19/Slovenia/6453/2020\|EPI_ISL_635228\|2020-03- | ···················· | ···················· | ···················· | ···················· | ···················· | ···················· |
| Q | 1384 hCoV-19/Slovenia/6455/2020\|EPI_ISL_635229\|2020-03- | ···················· | ···················· | ···················· | ···················· | ···················· | ···················· |
| Q | 1385 hCoV-19/Slovenia/6456/2020\|EPI_ISL_635230\|2020-03- | ···················· | ···················· | ···················· | ···················· | ···················· | ···················· |
| Q | 1386 hCoV-19/Slovenia/6462/2020\|EPI_ISL_635231\|2020-03- | ···················· | ···················· | ···················· | ···················· | ···················· | ···················· |
| Q | 1387 hCoV-19/Slovenia/6621/2020\|EPI_ISL_635232\|2020-03- | ···················· | ···················· | ···················· | ···················· | ···················· | ···················· |
| Q | 1388 hCoV-19/Slovenia/6667/2020\|EPI_ISL_635233\|2020-03- | ···················· | ···················· | ···················· | ···················· | ···················· | ···················· |
| Q | 1389 hCoV-19/Slovenia/6706/2020\|EPI_ISL_635234\|2020-03- | ···················· | ···················· | ···················· | ···················· | ···················· | ···················· |
| Q | 1390 hCoV-19/Slovenia/6999/2020\|EPI_ISL_635235\|2020-03- | ···················· | ···················· | ···················· | ···················· | ···················· | ···················· |
| Q | 1391 hCoV-19/Slovenia/7175/2020\|EPI_ISL_635236\|2020-03- | ···················· | ···················· | ···················· | ···················· | ···················· | ···················· |
| Q | 1392 hCoV-19/Slovenia/7213/2020\|EPI_ISL_635237\|2020-03- | ···················· | ···················· | ···················· | ···················· | ···················· | ···················· |
| Q | 1393 hCoV-19/Slovenia/7313/2020\|EPI_ISL_635238\|2020-03- | ···················· | ···················· | ···················· | ···················· | ···················· | ···················· |
| Q | 1394 hCoV-19/Slovenia/7360/2020\|EPI_ISL_635239\|2020-03- | ···················· | ···················· | ···················· | ···················· | ···················· | ···················· |
| Q | 1395 hCoV-19/Slovenia/7368/2020\|EPI_ISL_635240\|2020-03- | ···················· | ···················· | ···················· | ···················· | ···················· | ···················· |
| Q | 1396 hCoV-19/Slovenia/7455/2020\|EPI_ISL_635241\|2020-03- | ···················· | ···················· | ···················· | ···················· | ···················· | ···················· |
| Q | 1397 hCoV-19/Slovenia/7484/2020\|EPI_ISL_635242\|2020-03- | ···················· | ···················· | ···················· | ···················· | ···················· | ···················· |
| Q | 1398 hCoV-19/Slovenia/7540/2020\|EPI_ISL_635243\|2020-03- | ···················· | ···················· | ···················· | ···················· | ···················· | ···················· |
| Q | 1399 hCoV-19/Slovenia/7727/2020\|EPI_ISL_635244\|2020-03- | ···················· | ···················· | ···················· | ···················· | ···················· | ···················· |
| Q | 1400 hCoV-19/Slovenia/7735/2020\|EPI_ISL_635245\|2020-03- | ···················· | ···················· | ···················· | ···················· | ···················· | ···················· |
| Q | 1401 hCoV-19/Slovenia/7908/2020\|EPI_ISL_635246\|2020-03- | ···················· | ···················· | ···················· | ···················· | ···················· | ···················· |
| Q | 1402 hCoV-19/Slovenia/7990/2020\|EPI_ISL_635248\|2020-03- | ···················· | ···················· | ···················· | ···················· | ···················· | ···················· |
| Q | 1403 hCoV-19/Slovenia/8784/2020\|EPI_ISL_635250\|2020-03- | ···················· | ···················· | ···················· | ···················· | ···················· | ···················· |
| Q | 1404 hCoV-19/Slovenia/8852/2020\|EPI_ISL_635251\|2020-03- | ···················· | ···················· | ···················· | ···················· | ···················· | ···················· |
| Q | 1405 hCoV-19/Slovenia/10306/2020\|EPI_ISL_635252\|2020-03 | ···················· | ···················· | ···················· | ···················· | ···················· | ···················· |
| Q | 1407 hCoV-19/Slovenia/10906/2020\|EPI_ISL_635254\|2020-03 | ···················· | ···················· | ···················· | ···················· | ···················· | ···················· |
| Q | 1409 hCoV-19/Slovenia/11172/2020\|EPI_ISL_635256\|2020-03 | ···················· | ···················· | ···················· | ···················· | ···················· | ···················· |
| Q | 1411 hCoV-19/Slovenia/12230/2020\|EPI_ISL_635258\|2020-03 | ···················· | ···················· | ···················· | ···················· | ···················· | ···················· |

| | | | | | | | |
|---|---|---|---|---|---|---|---|
| Q | 1413 hCoV-19/Slovenia/12606/2020\|EPI_ISL_635260\|2020-03 | ···················· | ···················· | ···················· | ···················· | ···················· | ···················· |
| Q | 1414 hCoV-19/Slovenia/12692/2020\|EPI_ISL_635261\|2020-03 | ···················· | ···················· | ···················· | ···················· | ···················· | ···················· |
| Q | 1415 hCoV-19/Slovenia/12886/2020\|EPI_ISL_635262\|2020-03 | ···················· | ···················· | ···················· | ···················· | ···················· | ···················· |
| Q | 1417 hCoV-19/Slovenia/13052/2020\|EPI_ISL_635264\|2020-03 | ···················· | ···················· | ···················· | ···················· | ···················· | ···················· |
| Q | 1418 hCoV-19/Slovenia/13456/2020\|EPI_ISL_635265\|2020-03 | ···················· | ···················· | ···················· | ···················· | ···················· | ···················· |
| Q | 1419 hCoV-19/Slovenia/13487/2020\|EPI_ISL_635266\|2020-03 | ···················· | ···················· | ···················· | ···················· | ···················· | ···················· |
| Q | 1420 hCoV-19/Slovenia/V0-10433/2020\|EPI_ISL_635267\|2020 | ···················· | ···················· | ···················· | ···················· | ···················· | ···················· |
| Q | 1421 hCoV-19/Slovenia/V0-1288/2020\|EPI_ISL_635269\|2020- | ···················· | ···················· | ···················· | ···················· | ···················· | ···················· |
| Q | 1422 hCoV-19/Slovenia/V0-1421/2020\|EPI_ISL_635270\|2020- | ···················· | ···················· | ···················· | ···················· | ···················· | ···················· |
| Q | 1424 hCoV-19/Slovenia/V0-15921/2020\|EPI_ISL_635272\|2020 | ···················· | ···················· | ···················· | ···················· | ···················· | ···················· |
| Q | 1425 hCoV-19/Slovenia/V0-17284/2020\|EPI_ISL_635274\|2020 | ···················· | ···················· | ···················· | ···················· | ···················· | ···················· |
| Q | 1426 hCoV-19/Slovenia/V0-17285/2020\|EPI_ISL_635275\|2020 | ···················· | ···················· | ···················· | ···················· | ···················· | ···················· |
| Q | 1427 hCoV-19/Slovenia/V0-1731/2020\|EPI_ISL_635276\|2020- | ···················· | ···················· | ···················· | ···················· | ···················· | ···················· |
| Q | 1428 hCoV-19/Slovenia/V0-17818/2020\|EPI_ISL_635277\|2020 | ···················· | ···················· | ···················· | ···················· | ···················· | ···················· |
| Q | 1429 hCoV-19/Slovenia/V0-2250/2020\|EPI_ISL_635278\|2020- | ···················· | ···················· | ···················· | ···················· | ···················· | ···················· |
| Q | 1430 hCoV-19/Slovenia/V0-2804/2020\|EPI_ISL_635281\|2020- | ···················· | ···················· | ···················· | ···················· | ···················· | ···················· |
| Q | 1431 hCoV-19/Slovenia/V0-282/2020\|EPI_ISL_635282\|2020-0 | ···················· | ···················· | ···················· | ···················· | ···················· | ···················· |
| Q | 1432 hCoV-19/Slovenia/V0-2870/2020\|EPI_ISL_635283\|2020- | ···················· | ···················· | ···················· | ···················· | ···················· | ···················· |
| Q | 1433 hCoV-19/Slovenia/V0-2949/2020\|EPI_ISL_635284\|2020- | ···················· | ···················· | ···················· | ···················· | ···················· | ···················· |
| Q | 1434 hCoV-19/Slovenia/V0-3610/2020\|EPI_ISL_635285\|2020- | ···················· | ···················· | ···················· | ···················· | ···················· | ···················· |
| Q | 1435 hCoV-19/Slovenia/V0-3902/2020\|EPI_ISL_635286\|2020- | ···················· | ···················· | ···················· | ···················· | ···················· | ···················· |
| Q | 1436 hCoV-19/Slovenia/V0-408/2020\|EPI_ISL_635287\|2020-0 | ···················· | ···················· | ···················· | ···················· | ···················· | ···················· |
| Q | 1437 hCoV-19/Slovenia/V0-409/2020\|EPI_ISL_635288\|2020-0 | ···················· | ···················· | ···················· | ···················· | ···················· | ···················· |
| Q | 1438 hCoV-19/Slovenia/V0-4311/2020\|EPI_ISL_635289\|2020- | ···················· | ···················· | ···················· | ···················· | ···················· | ···················· |
| Q | 1439 hCoV-19/USA/CA-ALSR-2018/2020\|EPI_ISL_635308\|2020- | ···················· | ·A·················· | ···················· | ···················· | ···················· | ···················· |
| Q | 1440 hCoV-19/USA/CA-ALSR-2019/2020\|EPI_ISL_635309\|2020- | ···················· | ·A·················· | ···················· | ···················· | ···················· | ···················· |
| Q | 1441 hCoV-19/USA/CA-ALSR-2022/2020\|EPI_ISL_635310\|2020- | ···················· | ·A·················· | ···················· | ···················· | ···················· | ···················· |
| Q | 1442 hCoV-19/USA/CA-ALSR-2023/2020\|EPI_ISL_635311\|2020- | ···················· | ·A·················· | ···················· | ···················· | ···················· | ···················· |
| Q | 1443 hCoV-19/USA/CA-ALSR-2025/2020\|EPI_ISL_635312\|2020- | ···················· | ·A·················· | ···················· | ···················· | ···················· | ···················· |
| Q | 1444 hCoV-19/USA/CA-ALSR-2026/2020\|EPI_ISL_635313\|2020- | ···················· | ·A·················· | ···················· | ···················· | ···················· | ···················· |
| Q | 1445 hCoV-19/USA/CA-ALSR-2041/2020\|EPI_ISL_635314\|2020- | ···················· | ·A·················· | ···················· | ···················· | ···················· | ···················· |
| Q | 1446 hCoV-19/USA/CA-ALSR-2043/2020\|EPI_ISL_635315\|2020- | ···················· | ·A·················· | ···················· | ···················· | ···················· | ···················· |
| Q | 1447 hCoV-19/USA/CA-ALSR-2047/2020\|EPI_ISL_635316\|2020- | ···················· | ···················· | ···················· | ···················· | ···················· | ···················· |
| Q | 1448 hCoV-19/USA/CA-ALSR-2050/2020\|EPI_ISL_635317\|2020- | ···················· | ·A·················· | ···················· | ···················· | ···················· | ···················· |
| Q | 1449 hCoV-19/Netherlands/LI-RIVM-10313/2020\|EPI_ISL_636 | ···················· | ···················· | ···················· | ···················· | ···················· | ···················· |
| Q | 1450 hCoV-19/Netherlands/LI-RIVM-10330/2020\|EPI_ISL_636 | ···················· | ···················· | ···················· | ···················· | ···················· | ···················· |
| Q | 1451 hCoV-19/Netherlands/LI-RIVM-10331/2020\|EPI_ISL_636 | ···················· | ···················· | ···················· | ···················· | ···················· | ···················· |
| Q | 1452 hCoV-19/Netherlands/LI-RIVM-10335/2020\|EPI_ISL_636 | ···················· | ···················· | ···················· | ···················· | ···················· | ···················· |
| Q | 1453 hCoV-19/Netherlands/LI-RIVM-10347/2020\|EPI_ISL_636 | ···················· | ···················· | ···················· | ···················· | ···················· | ···················· |
| Q | 1454 hCoV-19/Netherlands/LI-RIVM-10354/2020\|EPI_ISL_636 | ···················· | ···················· | ···················· | ···················· | ···················· | ···················· |
| Q | 1455 hCoV-19/Netherlands/LI-RIVM-10381/2020\|EPI_ISL_636 | ···················· | ···················· | ···················· | ···················· | ······T············· | ···················· |
| Q | 1456 hCoV-19/Netherlands/NB-RIVM-10289/2020\|EPI_ISL_636 | ···················· | ···················· | ···················· | ···················· | ···················· | ···················· |
| Q | 1457 hCoV-19/Netherlands/NB-RIVM-10290/2020\|EPI_ISL_636 | ···················· | ···················· | ···················· | ···················· | ···················· | ···················· |
| Q | 1458 hCoV-19/Netherlands/NB-RIVM-10293/2020\|EPI_ISL_636 | ···················· | ···················· | ···················· | ···················· | ···················· | ···················· |
| Q | 1459 hCoV-19/Netherlands/NB-RIVM-10294/2020\|EPI_ISL_636 | ···················· | ···················· | ···················· | ···················· | ···················· | ···················· |
| Q | 1460 hCoV-19/Netherlands/NB-RIVM-10298/2020\|EPI_ISL_636 | ···················· | ···················· | ···················· | ···················· | ···················· | ···················· |
| Q | 1461 hCoV-19/Netherlands/NB-RIVM-10299/2020\|EPI_ISL_636 | ···················· | ···················· | ···················· | ···················· | ···················· | ···················· |
| Q | 1462 hCoV-19/Netherlands/NB-RIVM-10305/2020\|EPI_ISL_636 | ···················· | ···················· | ···················· | ···················· | ···················· | ···················· |
| Q | 1463 hCoV-19/Netherlands/NB-RIVM-10318/2020\|EPI_ISL_636 | ···················· | ···················· | ···················· | ···················· | ···················· | ···················· |
| Q | 1464 hCoV-19/Netherlands/NB-RIVM-10343/2020\|EPI_ISL_636 | ···················· | ···················· | ···················· | ···················· | ···················· | ···················· |
| Q | 1465 hCoV-19/Netherlands/NB-RIVM-10344/2020\|EPI_ISL_636 | ···················· | ···················· | ···················· | ···················· | ···················· | ···················· |
| Q | 1466 hCoV-19/Netherlands/NB-RIVM-10361/2020\|EPI_ISL_636 | ···················· | ···················· | ···················· | ···················· | ···················· | ···················· |
| Q | 1467 hCoV-19/Netherlands/NB-RIVM-10362/2020\|EPI_ISL_636 | ···················· | ···················· | ···················· | ···················· | ···················· | ···················· |
| Q | 1468 hCoV-19/Netherlands/NB-RIVM-10363/2020\|EPI_ISL_636 | ···················· | ···················· | ···················· | ···················· | ···················· | ···················· |
| Q | 1469 hCoV-19/Netherlands/NB-RIVM-10397/2020\|EPI_ISL_636 | ···················· | ···················· | ···················· | ···················· | ···················· | ···················· |
| Q | 1470 hCoV-19/Netherlands/NB-RIVM-10398/2020\|EPI_ISL_636 | ···················· | ·A·················· | ···················· | ···················· | ···················· | ···A················ |
| Q | 1471 hCoV-19/Netherlands/NH-RIVM-10291/2020\|EPI_ISL_636 | ···················· | ···················· | ···················· | ···················· | ···················· | ···················· |
| Q | 1472 hCoV-19/Netherlands/NH-RIVM-10312/2020\|EPI_ISL_636 | ···················· | ···················· | ···················· | ···················· | ···················· | ···················· |
| Q | 1473 hCoV-19/Netherlands/NH-RIVM-10315/2020\|EPI_ISL_636 | ···················· | ···················· | ···················· | ···················· | ···················· | ···················· |
| Q | 1474 hCoV-19/Netherlands/NH-RIVM-10341/2020\|EPI_ISL_636 | ···················· | ···················· | ···················· | ···················· | ···················· | ···················· |
| Q | 1475 hCoV-19/Netherlands/NH-RIVM-10342/2020\|EPI_ISL_636 | ···················· | ···················· | ···················· | ···················· | ···················· | ···················· |
| Q | 1476 hCoV-19/Netherlands/NH-RIVM-10353/2020\|EPI_ISL_636 | ···················· | ···················· | ···················· | ···················· | ···················· | ···················· |
| Q | 1477 hCoV-19/Netherlands/NH-RIVM-10357/2020\|EPI_ISL_636 | ···················· | ···················· | ···················· | ···················· | ···················· | ···················· |
| Q | 1478 hCoV-19/Netherlands/NH-RIVM-10387/2020\|EPI_ISL_636 | ···················· | ···················· | ···················· | ···················· | ···················· | ···················· |
| Q | 1479 hCoV-19/Netherlands/NH-RIVM-10394/2020\|EPI_ISL_636 | ···················· | ···················· | ···················· | ···················· | ······T············· | ···················· |
| Q | 1480 hCoV-19/Netherlands/NH-RIVM-10395/2020\|EPI_ISL_636 | ···················· | ···················· | ···················· | ···················· | ···················· | ···················· |
| Q | 1481 hCoV-19/Netherlands/NH-RIVM-10396/2020\|EPI_ISL_636 | ···················· | ···················· | ···················· | ···················· | ···················· | ···················· |
| Q | 1482 hCoV-19/Netherlands/OV-RIVM-10306/2020\|EPI_ISL_636 | ···················· | ···················· | ···················· | ···················· | ···················· | ···················· |
| Q | 1483 hCoV-19/Netherlands/UT-RIVM-10349/2020\|EPI_ISL_636 | ···················· | ···················· | ···················· | ···················· | ···················· | ···················· |
| Q | 1484 hCoV-19/Netherlands/UT-RIVM-10350/2020\|EPI_ISL_636 | ···················· | ···················· | ···················· | ···················· | ···················· | ···················· |
| Q | 1485 hCoV-19/Netherlands/UT-RIVM-10351/2020\|EPI_ISL_636 | ···················· | ···················· | ···················· | ···················· | ···················· | ···················· |
| Q | 1486 hCoV-19/Netherlands/UT-RIVM-10359/2020\|EPI_ISL_636 | ···················· | ···················· | ···················· | ···················· | ···················· | ···················· |
| Q | 1487 hCoV-19/Netherlands/UT-RIVM-10360/2020\|EPI_ISL_636 | ···················· | ···················· | ···················· | ···················· | ···················· | ···················· |
| Q | 1488 hCoV-19/Netherlands/UT-RIVM-10369/2020\|EPI_ISL_636 | ···················· | ···················· | ···················· | ···················· | ···················· | ···················· |
| Q | 1489 hCoV-19/Netherlands/UT-RIVM-10377/2020\|EPI_ISL_636 | ···················· | ···················· | ···················· | ···················· | ···················· | ···················· |
| Q | 1490 hCoV-19/Netherlands/UT-RIVM-10380/2020\|EPI_ISL_636 | ···················· | ···················· | ···················· | ···················· | ···················· | ···················· |
| Q | 1491 hCoV-19/Netherlands/UT-RIVM-10383/2020\|EPI_ISL_636 | ···················· | ···················· | ···················· | ···················· | ···················· | ···················· |
| Q | 1492 hCoV-19/Netherlands/UT-RIVM-10384/2020\|EPI_ISL_636 | ···················· | ···················· | ···················· | ···················· | ···················· | ···················· |
| Q | 1493 hCoV-19/Netherlands/UT-RIVM-10400/2020\|EPI_ISL_636 | ···················· | ···················· | ···················· | ···················· | ······T············· | ···················· |
| Q | 1494 hCoV-19/Netherlands/ZE-RIVM-10295/2020\|EPI_ISL_636 | ···················· | ···················· | ···················· | ···················· | ···················· | ···················· |

| | | | | | | | | |
|---|---|---|---|---|---|---|---|---|
| Q | 1495 | hCoV-19/Netherlands/ZE-RIVM-10296/2020\|EPI_ISL_636 | .................... | .................... | .................... | .................... | .................... | .................... |
| Q | 1496 | hCoV-19/Netherlands/ZE-RIVM-10352/2020\|EPI_ISL_636 | .................... | .................... | .................... | .................... | .................... | .................... |
| Q | 1497 | hCoV-19/Netherlands/ZE-RIVM-10355/2020\|EPI_ISL_636 | .................... | .................... | .................... | .................... | .................... | .................... |
| Q | 1498 | hCoV-19/Netherlands/ZE-RIVM-10356/2020\|EPI_ISL_636 | .................... | .................... | .................... | .................... | .................... | .................... |
| Q | 1499 | hCoV-19/Netherlands/ZE-RIVM-10375/2020\|EPI_ISL_636 | .................... | .................... | .................... | .................... | .................... | .................... |
| Q | 1500 | hCoV-19/Netherlands/ZE-RIVM-10376/2020\|EPI_ISL_636 | .................... | .................... | .................... | .................... | .................... | .................... |
| Q | 1501 | hCoV-19/Netherlands/ZH-RIVM-10297/2020\|EPI_ISL_636 | .................... | .................... | .................... | .................... | .................... | .................... |
| Q | 1502 | hCoV-19/Netherlands/ZH-RIVM-10310/2020\|EPI_ISL_636 | .................... | .................... | .................... | .................... | .................... | .................... |
| Q | 1503 | hCoV-19/Netherlands/ZH-RIVM-10316/2020\|EPI_ISL_636 | .................... | .................... | .................... | .................... | .................... | .................... |
| Q | 1504 | hCoV-19/Netherlands/ZH-RIVM-10333/2020\|EPI_ISL_636 | .................... | .................... | .................... | .................... | .................... | .................... |
| Q | 1505 | hCoV-19/Netherlands/ZH-RIVM-10334/2020\|EPI_ISL_636 | .................... | .................... | .................... | .................... | .................... | .................... |
| Q | 1506 | hCoV-19/Netherlands/ZH-RIVM-10339/2020\|EPI_ISL_636 | .................... | .................... | .................... | .................... | .................... | .................... |
| Q | 1507 | hCoV-19/Netherlands/ZH-RIVM-10340/2020\|EPI_ISL_636 | .................... | .................... | .................... | .................... | .................... | .................... |
| Q | 1508 | hCoV-19/Netherlands/ZH-RIVM-10348/2020\|EPI_ISL_636 | .................... | .................... | .................... | .................... | .................... | .................... |
| Q | 1509 | hCoV-19/Netherlands/ZH-RIVM-10358/2020\|EPI_ISL_636 | .................... | .................... | .................... | .................... | .................... | .................... |
| Q | 1510 | hCoV-19/St Eustatius/BQ-RIVM-10370/2020\|EPI_ISL_63 | .................... | .................... | .................... | .................... | .................... | .................... |
| Q | 1511 | hCoV-19/St Eustatius/BQ-RIVM-10371/2020\|EPI_ISL_63 | .................... | .................... | .................... | .................... | .................... | .................... |
| Q | 1512 | hCoV-19/St Eustatius/BQ-RIVM-10372/2020\|EPI_ISL_63 | .................... | .................... | .................... | .................... | .................... | .................... |
| Q | 1513 | hCoV-19/St Eustatius/BQ-RIVM-10373/2020\|EPI_ISL_63 | .................... | .................... | .................... | .................... | .................... | .................... |
| Q | 1514 | hCoV-19/Lithuania/MR-LUHS-Eilnr172/2020\|EPI_ISL_63 | .................... | .................... | .................... | .................... | .................... | .................... |
| Q | 1520 | hCoV-19/Belgium/ULG-10631/2020\|EPI_ISL_636610\|2020 | .................... | .................... | .................... | .................... | .................... | .................... |
| Q | 1522 | hCoV-19/Belgium/ULG-10633/2020\|EPI_ISL_636612\|2020 | .................... | .................... | .................... | .................... | .................... | .................... |
| Q | 1523 | hCoV-19/Belgium/ULG-10635/2020\|EPI_ISL_636613\|2020 | .................... | .................... | .................... | .................... | .................... | .................... |
| Q | 1525 | hCoV-19/Belgium/ULG-10637/2020\|EPI_ISL_636615\|2020 | .................... | .................... | .................... | .................... | .................... | .................... |
| Q | 1526 | hCoV-19/Belgium/ULG-10661/2020\|EPI_ISL_636638\|2020 | .................... | .................... | .................... | .................... | .................... | .................... |
| Q | 1527 | hCoV-19/Belgium/ULG-10662/2020\|EPI_ISL_636639\|2020 | .................... | .................... | .................... | .................... | .................... | .................... |
| Q | 1529 | hCoV-19/Belgium/ULG-10665/2020\|EPI_ISL_636641\|2020 | .................... | .................... | .................... | .................... | .................... | .................... |
| Q | 1530 | hCoV-19/Belgium/ULG-10666/2020\|EPI_ISL_636642\|2020 | .................... | .................... | .................... | .................... | .................... | .................... |
| Q | 1531 | hCoV-19/Belgium/ULG-10667/2020\|EPI_ISL_636643\|2020 | .................... | .................... | .................... | .................... | .................... | .................... |
| Q | 1535 | hCoV-19/Belgium/ULG-10672/2020\|EPI_ISL_636647\|2020 | .................... | .................... | .................... | .................... | .................... | .................... |
| Q | 1536 | hCoV-19/Belgium/ULG-10674/2020\|EPI_ISL_636648\|2020 | .................... | .................... | .................... | .................... | .................... | .................... |
| Q | 1537 | hCoV-19/Wales/PHWC-48447D/2020\|EPI_ISL_628006\|2020 | .................... | .................... | .................... | .................... | .................... | .................... |
| Q | 1538 | hCoV-19/Wales/PHWC-48448C/2020\|EPI_ISL_628007\|2020 | .................... | .................... | .................... | .................... | .................... | .................... |
| Q | 1540 | hCoV-19/Wales/PHWC-4844B9/2020\|EPI_ISL_628009\|2020 | .................... | .................... | .................... | .................... | .................... | .................... |
| Q | 1541 | hCoV-19/Wales/PHWC-4844E6/2020\|EPI_ISL_628010\|2020 | .................... | .................... | .................... | .................... | .................... | .................... |
| Q | 1542 | hCoV-19/Wales/PHWC-4844F5/2020\|EPI_ISL_628011\|2020 | .................... | .................... | .................... | .................... | .................... | .................... |
| Q | 1543 | hCoV-19/Wales/PHWC-484501/2020\|EPI_ISL_628012\|2020 | .................... | .................... | .................... | .................... | .................... | .................... |
| Q | 1544 | hCoV-19/Wales/PHWC-484510/2020\|EPI_ISL_628013\|2020 | .................... | .................... | .................... | .................... | .................... | .................... |
| Q | 1545 | hCoV-19/Wales/PHWC-48453E/2020\|EPI_ISL_628014\|2020 | .................... | .................... | .................... | .................... | .................... | .................... |
| Q | 1548 | hCoV-19/Wales/PHWC-484598/2020\|EPI_ISL_628017\|2020 | .................... | .................... | .................... | .................... | .................... | .................... |
| Q | 1549 | hCoV-19/Wales/PHWC-4845A7/2020\|EPI_ISL_628018\|2020 | .................... | .................... | .................... | .................... | .................... | .................... |
| Q | 1550 | hCoV-19/Wales/PHWC-4845B6/2020\|EPI_ISL_628019\|2020 | .................... | .................... | .................... | .................... | .................... | .................... |
| Q | 1552 | hCoV-19/Wales/PHWC-484765/2020\|EPI_ISL_628041\|2020 | .................... | .................... | .................... | .................... | .................... | .................... |
| Q | 1553 | hCoV-19/Wales/PHWC-484783/2020\|EPI_ISL_628042\|2020 | .................... | .................... | .................... | .................... | .................... | .................... |
| Q | 1554 | hCoV-19/Wales/PHWC-4847B0/2020\|EPI_ISL_628043\|2020 | .................... | .................... | .................... | .................... | .................... | .................... |
| Q | 1555 | hCoV-19/Wales/PHWC-4847CF/2020\|EPI_ISL_628044\|2020 | .................... | .................... | .................... | .................... | .................... | .................... |
| Q | 1556 | hCoV-19/Wales/PHWC-4847DE/2020\|EPI_ISL_628045\|2020 | .................... | .................... | .................... | .................... | .................... | .................... |
| Q | 1557 | hCoV-19/Wales/PHWC-4847ED/2020\|EPI_ISL_628046\|2020 | .................... | .................... | .................... | .................... | .................... | .................... |
| Q | 1558 | hCoV-19/Wales/PHWC-484808/2020\|EPI_ISL_628047\|2020 | .................... | .................... | .................... | .................... | .................... | .................... |
| Q | 1559 | hCoV-19/Wales/PHWC-484817/2020\|EPI_ISL_628048\|2020 | .................... | .................... | .................... | .................... | .................... | .................... |
| Q | 1560 | hCoV-19/Wales/PHWC-484826/2020\|EPI_ISL_628049\|2020 | .................... | .................... | .................... | .................... | .................... | .................... |
| Q | 1561 | hCoV-19/Wales/PHWC-484835/2020\|EPI_ISL_628050\|2020 | .................... | .................... | .................... | .................... | .................... | .................... |
| Q | 1562 | hCoV-19/Wales/PHWC-484853/2020\|EPI_ISL_628051\|2020 | .................... | .................... | .................... | .................... | .................... | .................... |
| Q | 1563 | hCoV-19/Wales/PHWC-484871/2020\|EPI_ISL_628052\|2020 | .................... | .................... | .................... | .................... | .................... | .................... |
| Q | 1564 | hCoV-19/Wales/PHWC-484880/2020\|EPI_ISL_628053\|2020 | .................... | .................... | .................... | .................... | .................... | .................... |
| Q | 1565 | hCoV-19/Wales/PHWC-48489F/2020\|EPI_ISL_628054\|2020 | .................... | .................... | .................... | .................... | .................... | .................... |
| Q | 1566 | hCoV-19/Wales/PHWC-4848CC/2020\|EPI_ISL_628055\|2020 | .................... | .................... | .................... | .................... | .................... | .................... |
| Q | 1567 | hCoV-19/Wales/PHWC-4848EA/2020\|EPI_ISL_628056\|2020 | .................... | .................... | .................... | .................... | .................... | .................... |
| Q | 1568 | hCoV-19/Wales/PHWC-4848F9/2020\|EPI_ISL_628057\|2020 | .................... | .................... | .................... | .................... | .................... | .................... |
| Q | 1569 | hCoV-19/Wales/PHWC-484914/2020\|EPI_ISL_628058\|2020 | .................... | .................... | .................... | .................... | .................... | .................... |
| Q | 1570 | hCoV-19/Wales/PHWC-484923/2020\|EPI_ISL_628059\|2020 | .................... | .................... | .................... | .................... | .NN................. | .................... |
| Q | 1571 | hCoV-19/Wales/PHWC-484932/2020\|EPI_ISL_628060\|2020 | .................... | .................... | .................... | .................... | .................... | .................... |
| Q | 1573 | hCoV-19/Wales/PHWC-48496F/2020\|EPI_ISL_628062\|2020 | .................... | .................... | .................... | .................... | .................... | .................... |
| Q | 1574 | hCoV-19/Wales/PHWC-48497E/2020\|EPI_ISL_628063\|2020 | .................... | .................... | .................... | .................... | .................... | .................... |
| Q | 1575 | hCoV-19/Wales/PHWC-48498D/2020\|EPI_ISL_628064\|2020 | .................... | .................... | .................... | .................... | .................... | .................... |
| Q | 1576 | hCoV-19/Wales/PHWC-4849AB/2020\|EPI_ISL_628065\|2020 | .................... | .................... | .................... | .................... | .................... | .................... |
| Q | 1577 | hCoV-19/Wales/PHWC-4849BA/2020\|EPI_ISL_628066\|2020 | .................... | .................... | .................... | .................... | .................... | .................... |
| Q | 1578 | hCoV-19/Wales/PHWC-4849F6/2020\|EPI_ISL_628067\|2020 | .................... | .................... | .................... | .................... | .................... | .................... |
| Q | 1579 | hCoV-19/Wales/PHWC-484A11/2020\|EPI_ISL_628068\|2020 | .................... | .................... | .................... | .................... | .................... | .................... |
| Q | 1580 | hCoV-19/Wales/PHWC-484A3F/2020\|EPI_ISL_628069\|2020 | .................... | .................... | .................... | .................... | .................... | .................... |
| Q | 1581 | hCoV-19/Wales/PHWC-484A4E/2020\|EPI_ISL_628070\|2020 | .................... | .................... | .................... | .................... | .................... | .................... |
| Q | 1582 | hCoV-19/Wales/PHWC-484A6C/2020\|EPI_ISL_628071\|2020 | .................... | .................... | .................... | .................... | .................... | .................... |
| Q | 1583 | hCoV-19/Wales/PHWC-484A7B/2020\|EPI_ISL_628072\|2020 | .................... | .................... | .................... | .................... | .................... | .................... |
| Q | 1584 | hCoV-19/Wales/PHWC-484A8A/2020\|EPI_ISL_628073\|2020 | .................... | .................... | .................... | .................... | .................... | .................... |
| Q | 1585 | hCoV-19/Wales/PHWC-484A99/2020\|EPI_ISL_628074\|2020 | .................... | .................... | .................... | .................... | .................... | .................... |
| Q | 1586 | hCoV-19/Wales/PHWC-484AA8/2020\|EPI_ISL_628075\|2020 | .................... | .................... | .................... | .................... | .................... | .................... |
| Q | 1588 | hCoV-19/Wales/PHWC-484AD5/2020\|EPI_ISL_628077\|2020 | .................... | .................... | .................... | .................... | .................... | .................... |
| Q | 1589 | hCoV-19/Wales/PHWC-484AE4/2020\|EPI_ISL_628078\|2020 | .................... | .................... | .................... | .................... | .................... | .................... |
| Q | 1590 | hCoV-19/Wales/PHWC-484AF3/2020\|EPI_ISL_628079\|2020 | .................... | .................... | .................... | .................... | .................... | .................... |
| Q | 1591 | hCoV-19/Wales/PHWC-484D09/2020\|EPI_ISL_628100\|2020 | .................... | .................... | .................... | .................... | .................... | .................... |

Q 1592 hCoV-19/Wales/PHWC-484D18/2020|EPI_ISL_628101|2020 ···················· ···················· ···················· ···················· ···················· ····················
Q 1594 hCoV-19/Wales/PHWC-484D36/2020|EPI_ISL_628103|2020 ···················· ···················· ···················· ···················· ···················· ····················
Q 1595 hCoV-19/Wales/PHWC-484D45/2020|EPI_ISL_628104|2020 ···················· ···················· ···················· ···················· ···················· ····················
Q 1596 hCoV-19/Wales/PHWC-484D54/2020|EPI_ISL_628105|2020 ···················· ···················· ···················· ···················· ···················· ····················
Q 1597 hCoV-19/Wales/PHWC-484D63/2020|EPI_ISL_628106|2020 ···················· ···················· ···················· ···················· ···················· ····················
Q 1598 hCoV-19/Wales/PHWC-484D72/2020|EPI_ISL_628107|2020 ···················· ···················· ···················· ···················· ···················· ····················
Q 1599 hCoV-19/Wales/PHWC-484D81/2020|EPI_ISL_628108|2020 ···················· ···················· ···················· ···················· ···················· ····················
Q 1600 hCoV-19/Wales/PHWC-484D90/2020|EPI_ISL_628109|2020 ···················· ···················· ···················· ···················· ···················· ····················
Q 1601 hCoV-19/Wales/PHWC-484DDC/2020|EPI_ISL_628112|2020 ···················· ···················· ···················· ···················· ···················· ····················
Q 1602 hCoV-19/Wales/PHWC-484DEB/2020|EPI_ISL_628113|2020 ···················· ···················· ···················· ···················· ···················· ····················
Q 1603 hCoV-19/Wales/PHWC-484DFA/2020|EPI_ISL_628114|2020 ···················· ···················· ···················· ···················· ···················· ····················
Q 1604 hCoV-19/Wales/PHWC-484E06/2020|EPI_ISL_628115|2020 ···················· ···················· ···················· ···················· ···················· ····················
Q 1605 hCoV-19/England/MILK-ACC08E/2020|EPI_ISL_629338|20 ···················· ···················· ···················· ···················· ···················· ····················
Q 1606 hCoV-19/England/MILK-ACC3FE/2020|EPI_ISL_629339|20 ···················· ···················· ···················· ···················· ···················· ····················
Q 1607 hCoV-19/England/MILK-AC9555/2020|EPI_ISL_629340|20 ···················· ···················· ···················· ···················· ···················· ····················
Q 1608 hCoV-19/England/MILK-ACBE94/2020|EPI_ISL_629341|20 ···················· ···················· ···················· ···················· ···················· ····················
Q 1609 hCoV-19/England/MILK-ACEC79/2020|EPI_ISL_629342|20 ···················· ···················· ···················· ···················· ······T············· ····················
Q 1610 hCoV-19/England/MILK-ACEC1F/2020|EPI_ISL_629343|20 ···················· ···················· ···················· ···················· ···················· ····················
Q 1611 hCoV-19/England/MILK-AC8477/2020|EPI_ISL_629344|20 ···················· ···················· ···················· ···················· ···················· ····················
Q 1612 hCoV-19/England/MILK-ACE408/2020|EPI_ISL_629345|20 ···················· ···················· ···················· ···················· ···················· ····················
Q 1613 hCoV-19/England/MILK-ACBC8B/2020|EPI_ISL_629346|20 ···················· ···················· ···················· ···················· ···················· ····················
Q 1614 hCoV-19/England/MILK-ACE444/2020|EPI_ISL_629348|20 ···················· ···················· ···················· ···················· ···················· ····················
Q 1615 hCoV-19/England/MILK-ACBF55/2020|EPI_ISL_629349|20 ···················· ···················· ···················· ···················· ···················· ····················
Q 1616 hCoV-19/England/MILK-ACE945/2020|EPI_ISL_629370|20 ···················· ···················· ···················· ···················· ···················· ····················
Q 1617 hCoV-19/England/MILK-AC836B/2020|EPI_ISL_629371|20 ···················· ···················· ···················· ···················· ····T··············· ····················
Q 1618 hCoV-19/England/MILK-AC7D9B/2020|EPI_ISL_629372|20 ···················· ···················· ···················· ···················· ···················· ····················
Q 1619 hCoV-19/England/MILK-AC9643/2020|EPI_ISL_629373|20 ···················· ···················· ···················· ···················· ···················· ····················
Q 1620 hCoV-19/England/MILK-ACBF64/2020|EPI_ISL_629374|20 ···················· ···················· ···················· ···················· ···················· ····················
Q 1621 hCoV-19/England/MILK-AC8486/2020|EPI_ISL_629375|20 ···················· ···················· ···················· ···················· ···················· ····················
Q 1622 hCoV-19/England/MILK-ACBD4C/2020|EPI_ISL_629376|20 ···················· ···················· ···················· ···················· ···················· ····················
Q 1623 hCoV-19/England/MILK-ACC07F/2020|EPI_ISL_629377|20 ···················· ···················· ···················· ···················· ···················· ····················
Q 1624 hCoV-19/England/MILK-AC8592/2020|EPI_ISL_629378|20 ···················· ···················· ···················· ···················· ···················· ····················
Q 1625 hCoV-19/England/MILK-ACED67/2020|EPI_ISL_629379|20 ···················· ···················· ···················· ···················· ···················· ····················
Q 1626 hCoV-19/England/MILK-ACE295/2020|EPI_ISL_629380|20 ···················· ···················· ···················· ···················· ···················· ····················
Q 1627 hCoV-19/England/MILK-ACC3EF/2020|EPI_ISL_629381|20 ···················· ···················· ···················· ···················· ···················· ····················
Q 1628 hCoV-19/England/MILK-ACBF0A/2020|EPI_ISL_629382|20 ···················· ···················· ···················· ···················· ···················· ····················
Q 1629 hCoV-19/England/MILK-ACE4AE/2020|EPI_ISL_629383|20 ···················· ···················· ···················· ···················· ···················· ····················
Q 1630 hCoV-19/England/MILK-AC8583/2020|EPI_ISL_629384|20 ···················· ···················· ···················· ···················· ···················· ····················
Q 1631 hCoV-19/England/MILK-ACC8A4/2020|EPI_ISL_629385|20 ···················· ·A·················· ···················· ···················· ···················· ···A················
Q 1632 hCoV-19/England/MILK-ACC279/2020|EPI_ISL_629386|20 ···················· ···················· ···················· ···················· ···················· ····················
Q 1633 hCoV-19/England/MILK-AC8662/2020|EPI_ISL_629387|20 ···················· ···················· ···················· ···················· ···················· ····················
Q 1634 hCoV-19/England/MILK-ACC3D0/2020|EPI_ISL_629388|20 ···················· ·A·················· ···················· ···················· ···················· ···A················
Q 1635 hCoV-19/England/MILK-ACC74D/2020|EPI_ISL_629389|20 ···················· ···················· ···················· ···················· ······T············· ····················
Q 1636 hCoV-19/England/MILK-ACE383/2020|EPI_ISL_629390|20 ···················· ···················· ···················· ···················· ···················· ····················
Q 1637 hCoV-19/England/MILK-AC841D/2020|EPI_ISL_629391|20 ···················· ···················· ···················· ···················· ···················· ····················
Q 1638 hCoV-19/England/MILK-ACE8B1/2020|EPI_ISL_629392|20 ···················· ···················· ···················· ···················· ···················· ····················
Q 1639 hCoV-19/England/MILK-AC83C5/2020|EPI_ISL_629393|20 ···················· ···················· ···················· ···················· ···················· ····················
Q 1640 hCoV-19/England/MILK-AC8398/2020|EPI_ISL_629394|20 ···················· ···················· ···················· ···················· ···················· ····················
Q 1641 hCoV-19/England/MILK-ACBDF1/2020|EPI_ISL_629395|20 ···················· ···················· ···················· ···················· ···················· ····················
Q 1642 hCoV-19/England/MILK-AC94FE/2020|EPI_ISL_629396|20 ···················· ···················· ···················· ···················· ···················· ····················
Q 1643 hCoV-19/England/MILK-AC8310/2020|EPI_ISL_629397|20 ···················· ···················· ···················· ···················· ···················· ····················
Q 1644 hCoV-19/England/MILK-ACC2B5/2020|EPI_ISL_629399|20 ···················· ···················· ···················· ···················· ···················· ····················
Q 1645 hCoV-19/England/MILK-ACC895/2020|EPI_ISL_629400|20 ···················· ···················· ···················· ···················· ···················· ····················
Q 1646 hCoV-19/England/MILK-ACC0AC/2020|EPI_ISL_629401|20 ···················· ···················· ···················· ···················· ···················· ····················
Q 1647 hCoV-19/England/MILK-AC926D/2020|EPI_ISL_629402|20 ···················· ···················· ···················· ···················· ···················· ····················
Q 1648 hCoV-19/England/MILK-ACBE49/2020|EPI_ISL_629405|20 ···················· ···················· ···················· ···················· ···················· ····················
Q 1649 hCoV-19/England/MILK-ACBD6A/2020|EPI_ISL_629406|20 ···················· ···················· ···················· ···················· ···················· ····················
Q 1650 hCoV-19/England/MILK-AC7CBC/2020|EPI_ISL_629407|20 ···················· ···················· ···················· ···················· ···················· ····················
Q 1651 hCoV-19/England/MILK-ACC288/2020|EPI_ISL_629430|20 ···················· ···················· ···················· ···················· ······T············· ····················
Q 1652 hCoV-19/England/MILK-ACEDA3/2020|EPI_ISL_629431|20 ···················· ···················· ···················· ···················· ···················· ····················
Q 1653 hCoV-19/England/MILK-ACEC5B/2020|EPI_ISL_629432|20 ···················· ·A·················· ···················· ···················· ···················· ····················
Q 1654 hCoV-19/England/MILK-ACEBF4/2020|EPI_ISL_629433|20 ···················· ···················· ···················· ···················· ···················· ····················
Q 1655 hCoV-19/England/MILK-AC83E3/2020|EPI_ISL_629434|20 ···················· ···················· ···················· ···················· ···················· ····················
Q 1656 hCoV-19/England/MILK-AC8DC7/2020|EPI_ISL_629435|20 ···················· ···················· ···················· ···················· ······T············· ····················
Q 1657 hCoV-19/England/MILK-ACE6D5/2020|EPI_ISL_629436|20 ···················· ···················· ···················· ···················· ······T············· ····················
Q 1658 hCoV-19/England/MILK-AC8A2A/2020|EPI_ISL_629437|20 ···················· ···················· ···················· ···················· ···················· ····················
Q 1659 hCoV-19/England/MILK-ACF407/2020|EPI_ISL_629439|20 ···················· ···················· ···················· ···················· ···················· ····················
Q 1660 hCoV-19/England/MILK-ACE2A4/2020|EPI_ISL_629443|20 ···················· ···················· ···················· ···················· ···················· ····················
Q 1661 hCoV-19/England/MILK-AC84D1/2020|EPI_ISL_629444|20 ···················· ···················· ···················· ···················· ···················· ····················
Q 1662 hCoV-19/England/MILK-ACF197/2020|EPI_ISL_629445|20 ···················· ···················· ···················· ···················· ···················· ····················
Q 1663 hCoV-19/England/MILK-ACE040/2020|EPI_ISL_630317|20 ···················· ·A·················· ···················· ···················· ···················· ····················
Q 1664 hCoV-19/England/MILK-AC72D8/2020|EPI_ISL_630318|20 ···················· ···················· ···················· ···················· ···················· ····················
Q 1665 hCoV-19/England/MILK-ACDEA1/2020|EPI_ISL_630320|20 ···················· ···················· ···················· ···················· ······T············· ····················
Q 1666 hCoV-19/England/MILK-AC935B/2020|EPI_ISL_630321|20 ···················· ···················· ···················· ···················· ······T············· ····················
Q 1667 hCoV-19/England/MILK-ACDBE6/2020|EPI_ISL_630322|20 ···················· ···················· ···················· ···················· ···················· ····················
Q 1668 hCoV-19/England/MILK-AC942B/2020|EPI_ISL_630323|20 ···················· ···················· ···················· ···················· ···················· ····················
Q 1669 hCoV-19/England/MILK-ACCCC6/2020|EPI_ISL_630324|20 ···················· ···················· ···················· ···················· ···················· ····················
Q 1670 hCoV-19/England/MILK-ACDCA7/2020|EPI_ISL_630325|20 ···················· ···················· ···················· ···················· ···················· ····················
Q 1671 hCoV-19/England/MILK-AC7302/2020|EPI_ISL_630326|20 ···················· ···················· ···················· ···················· ···················· ····················
Q 1672 hCoV-19/England/MILK-AC7478/2020|EPI_ISL_630327|20 ···················· ···················· ···················· ···················· ···················· ····················

```
Q 1673 hCoV-19/England/MILK-AC73C6/2020|EPI_ISL_630328|20 ···················· ···················· ····················· ···················· ···················· ····················
Q 1674 hCoV-19/England/MILK-ACD050/2020|EPI_ISL_630329|20 ···················· ···················· ····················· ···················· ···················· ····················
Q 1675 hCoV-19/England/MILK-ACDC01/2020|EPI_ISL_630330|20 ···················· ···················· ····················· ···················· ···················· ····················
Q 1676 hCoV-19/England/MILK-ACE0F5/2020|EPI_ISL_630331|20 ···················· ···················· ····················· ···················· ···················· ····················
Q 1677 hCoV-19/England/MILK-ACDC2F/2020|EPI_ISL_630333|20 ···················· ···················· ····················· ···················· ···················· ····················
Q 1678 hCoV-19/England/MILK-ACD858/2020|EPI_ISL_630334|20 ···················· ···················· ····················· ···················· ···················· ····················
Q 1679 hCoV-19/England/ALDP-A66912/2020|EPI_ISL_630357|20 ···················· ···················· ····················· ···················· ···················· ····················
Q 1680 hCoV-19/England/ALDP-A6786F/2020|EPI_ISL_630358|20 ···················· ···················· ····················· ···················· ···················· ····················
Q 1681 hCoV-19/England/ALDP-A675E0/2020|EPI_ISL_630359|20 ···················· ···················· ····················· ···················· ···················· ····················
Q 1682 hCoV-19/England/ALDP-A67191/2020|EPI_ISL_630360|20 ···················· ···················· ····················· ···················· ···················· ····················
Q 1683 hCoV-19/England/ALDP-A6789C/2020|EPI_ISL_630361|20 ···················· ···················· ····················· ···················· ···················· ····················
Q 1684 hCoV-19/England/ALDP-A66ED7/2020|EPI_ISL_630362|20 ···················· ···················· ····················· ···················· ···················· ····················
Q 1685 hCoV-19/England/ALDP-A66E04/2020|EPI_ISL_630363|20 ···················· ···················· ····················· ···················· ···················· ····················
Q 1686 hCoV-19/England/ALDP-A67780/2020|EPI_ISL_630364|20 ···················· ···················· ····················· ···················· ···················· ····················
Q 1687 hCoV-19/England/ALDP-A67173/2020|EPI_ISL_630365|20 ···················· ···················· ····················· ···················· ···················· ····················
Q 1688 hCoV-19/England/ALDP-A67CAF/2020|EPI_ISL_630366|20 ···················· ···················· ····················· ···················· ···················· ····················
Q 1689 hCoV-19/England/ALDP-A679F3/2020|EPI_ISL_630367|20 ···················· ·A·················· ····················· ···················· ···················· ···A················
Q 1690 hCoV-19/England/ALDP-A67CCD/2020|EPI_ISL_630368|20 ···················· ···················· ····················· ···················· ···················· ····················
Q 1691 hCoV-19/England/ALDP-A672DA/2020|EPI_ISL_630369|20 ···················· ···················· ····················· ···················· ···················· ····················
Q 1692 hCoV-19/England/ALDP-A66BEF/2020|EPI_ISL_630370|20 ···················· ···················· ····················· ···················· ···················· ····················
Q 1693 hCoV-19/England/ALDP-A67A87/2020|EPI_ISL_630371|20 ···················· ···················· ····················· ···················· ···················· ····················
Q 1694 hCoV-19/England/ALDP-A686FB/2020|EPI_ISL_630372|20 ···················· ···················· ····················· ···················· ···················· ····················
Q 1695 hCoV-19/England/ALDP-A67735/2020|EPI_ISL_630373|20 ···················· ···················· ····················· ···················· ···················· ····················
Q 1696 hCoV-19/England/ALDP-A66D25/2020|EPI_ISL_630374|20 ···················· ···················· ····················· ···················· ···················· ····················
Q 1697 hCoV-19/England/ALDP-A676CF/2020|EPI_ISL_630375|20 ···················· ···················· ····················· ···················· ···················· ····················
Q 1698 hCoV-19/England/ALDP-A67A0F/2020|EPI_ISL_630376|20 ···················· ···················· ····················· ···················· ···················· ····················
Q 1699 hCoV-19/England/ALDP-A67A2D/2020|EPI_ISL_630377|20 ···················· ···················· ····················· ···················· ···················· ····················
Q 1700 hCoV-19/England/ALDP-A66B94/2020|EPI_ISL_630378|20 ···················· ···················· ····················· ···················· ···················· ····················
Q 1701 hCoV-19/England/ALDP-A67665/2020|EPI_ISL_630379|20 ···················· ·A·················· ····················· ···················· ···················· ····················
Q 1702 hCoV-19/England/ALDP-A685FE/2020|EPI_ISL_630380|20 ···················· ···················· ····················· ···················· ···················· ····················
Q 1703 hCoV-19/England/ALDP-A686DD/2020|EPI_ISL_630381|20 ···················· ···················· ····················· ···················· ···················· ····················
Q 1704 hCoV-19/England/ALDP-A68734/2020|EPI_ISL_630382|20 ···················· ···················· ····················· ···················· ···················· ····················
Q 1705 hCoV-19/England/ALDP-A675FF/2020|EPI_ISL_630383|20 ···················· ···················· ····················· ···················· ···················· ····················
Q 1706 hCoV-19/England/ALDP-A67AD2/2020|EPI_ISL_630384|20 ···················· ···················· ····················· ···················· ···················· ····················
Q 1707 hCoV-19/England/ALDP-A689E3/2020|EPI_ISL_630385|20 ···················· ···················· ····················· ···················· ···················· ····················
Q 1708 hCoV-19/England/ALDP-A66B85/2020|EPI_ISL_630386|20 ···················· ···················· ····················· ···················· ···················· ····················
Q 1709 hCoV-19/England/ALDP-A67CDC/2020|EPI_ISL_630387|20 ···················· ···················· ····················· ···················· ···················· ····················
Q 1710 hCoV-19/England/ALDP-A672E9/2020|EPI_ISL_630388|20 ···················· ···················· ····················· ···················· ···················· ····················
Q 1711 hCoV-19/England/ALDP-A67762/2020|EPI_ISL_630389|20 ···················· ···················· ····················· ···················· ···················· ····················
Q 1712 hCoV-19/England/ALDP-A66B2B/2020|EPI_ISL_630390|20 ···················· ···················· ····················· ···················· ···················· ····················
Q 1713 hCoV-19/England/ALDP-A67577/2020|EPI_ISL_630391|20 ···················· ···················· ····················· ···················· ···················· ····················
Q 1714 hCoV-19/England/ALDP-A689C5/2020|EPI_ISL_630392|20 ···················· ···················· ····················· ···················· ···················· ····················
Q 1715 hCoV-19/England/ALDP-A67146/2020|EPI_ISL_630393|20 ···················· ···················· ····················· ···················· ···················· ····················
Q 1716 hCoV-19/England/ALDP-A671CE/2020|EPI_ISL_630394|20 ···················· ···················· ····················· ···················· ···················· ····················
Q 1717 hCoV-19/England/ALDP-A66AA6/2020|EPI_ISL_630395|20 ···················· ···················· ····················· ···················· ···················· ····················
Q 1718 hCoV-19/England/ALDP-A66F3E/2020|EPI_ISL_630396|20 ···················· ···················· ····················· ···················· ···················· ····················
Q 1719 hCoV-19/England/ALDP-A66F5C/2020|EPI_ISL_630398|20 ···················· ···················· ····················· ···················· ···················· ····················
Q 1720 hCoV-19/England/ALDP-A67568/2020|EPI_ISL_630399|20 ···················· ···················· ····················· ···················· ···················· ····················
Q 1721 hCoV-19/England/ALDP-A66D43/2020|EPI_ISL_630412|20 ···················· ···················· ····················· ···················· ···················· ····················
Q 1722 hCoV-19/England/ALDP-A671FB/2020|EPI_ISL_630413|20 ···················· ···················· ····················· ···················· ···················· ····················
Q 1723 hCoV-19/England/ALDP-A67234/2020|EPI_ISL_630414|20 ···················· ·A·················· ····················· ···················· ···················· ···A················
Q 1724 hCoV-19/England/ALDP-A6798A/2020|EPI_ISL_630415|20 ···················· ···················· ····················· ···················· ···················· ····················
Q 1725 hCoV-19/England/ALDP-A68637/2020|EPI_ISL_630416|20 ···················· ···················· ····················· ···················· ···················· ····················
Q 1726 hCoV-19/England/ALDP-A6753B/2020|EPI_ISL_630417|20 ···················· ···················· ····················· ···················· ···················· ····················
Q 1727 hCoV-19/England/ALDP-A67A69/2020|EPI_ISL_630418|20 ···················· ···················· ····················· ···················· ···················· ····················
Q 1728 hCoV-19/England/ALDP-A6698B/2020|EPI_ISL_630419|20 ···················· ···················· ····················· ···················· ···················· ····················
Q 1729 hCoV-19/England/ALDP-A67C36/2020|EPI_ISL_630421|20 ···················· ···················· ····················· ···················· ···················· ····················
Q 1730 hCoV-19/England/ALDP-A66D16/2020|EPI_ISL_630422|20 ···················· ···················· ····················· ···················· ···················· ····················
Q 1731 hCoV-19/USA/TX-DSHS-1198/2020|EPI_ISL_631660|2020- ···················· ···················· ····················· ···················· ···················· ····················
Q 1732 hCoV-19/USA/TX-DSHS-1199/2020|EPI_ISL_631661|2020- ···················· ···················· ····················· ···················· ···················· ····················
Q 1733 hCoV-19/USA/NY-NYCPHL-000152/2020|EPI_ISL_631688|2 ···················· ···················· ····················· ···················· ···················· ····················
Q 1734 hCoV-19/USA/NY-NYCPHL-000153/2020|EPI_ISL_631689|2 ···················· ···················· ····················· ···················· ···················· ····················
Q 1735 hCoV-19/USA/NY-NYCPHL-000154/2020|EPI_ISL_631690|2 ···················· ···················· ····················· ···················· ···················· ····················
Q 1736 hCoV-19/USA/NY-NYCPHL-000157/2020|EPI_ISL_631693|2 ···················· ···················· ····················· ···················· ···················· ····················
Q 1737 hCoV-19/USA/NY-NYCPHL-000158/2020|EPI_ISL_631694|2 ···················· ···················· ····················· ···················· ···················· ····················
Q 1738 hCoV-19/USA/NY-NYCPHL-000159/2020|EPI_ISL_631695|2 ···················· ···················· ····················· ···················· ···················· ····················
Q 1739 hCoV-19/USA/NY-NYCPHL-000161/2020|EPI_ISL_631697|2 ···················· ···················· ····················· ···················· ···················· ····················
Q 1740 hCoV-19/USA/NY-NYCPHL-000162/2020|EPI_ISL_631698|2 ···················· ···················· ····················· ···················· ···················· ····················
Q 1741 hCoV-19/USA/NY-NYCPHL-000165/2020|EPI_ISL_631701|2 ···················· ···················· ····················· ···················· ···················· ····················
Q 1742 hCoV-19/USA/NY-NYCPHL-000166/2020|EPI_ISL_631702|2 ···················· ···················· ····················· ···················· ···················· ····················
Q 1743 hCoV-19/USA/NY-NYCPHL-000167/2020|EPI_ISL_631703|2 ···················· ···················· ····················· ···················· ···················· ····················
Q 1744 hCoV-19/USA/NY-NYCPHL-000168/2020|EPI_ISL_631704|2 ···················· ···················· ····················· ···················· ···················· ····················
Q 1745 hCoV-19/USA/NY-NYCPHL-000169/2020|EPI_ISL_631705|2 ···················· ···················· ····················· ···················· ···················· ····················
Q 1746 hCoV-19/USA/NY-NYCPHL-000170/2020|EPI_ISL_631706|2 ···················· ···················· ····················· ···················· ···················· ····················
Q 1747 hCoV-19/USA/NY-NYCPHL-000173/2020|EPI_ISL_631709|2 ···················· ···················· ····················· ···················· ···················· ····················
Q 1748 hCoV-19/USA/NY-NYCPHL-000174/2020|EPI_ISL_631710|2 ···················· ···················· ····················· ···················· ···················· ····················
Q 1749 hCoV-19/USA/NY-NYCPHL-000175/2020|EPI_ISL_631711|2 ···················· ···················· ····················· ···················· ···················· ····················
Q 1750 hCoV-19/USA/NY-NYCPHL-000177/2020|EPI_ISL_631712|2 ···················· ···················· ····················· ···················· ···················· ····················
Q 1751 hCoV-19/USA/NY-NYCPHL-000178/2020|EPI_ISL_631713|2 ···················· ···················· ····················· ···················· ···················· ····················
Q 1752 hCoV-19/USA/NY-NYCPHL-000181/2020|EPI_ISL_631716|2 ···················· ···················· ····················· ···················· ···················· ····················
```

| | | | | | | | |
|---|---|---|---|---|---|---|---|
| Q | 1753 hCoV-19/USA/NY-NYCPHL-000182/2020\|EPI_ISL_631717\|2 | ···················· | ···················· | ···················· | ···················· | ···················· | ···················· |
| Q | 1754 hCoV-19/USA/NY-NYCPHL-000183/2020\|EPI_ISL_631718\|2 | ···················· | ···················· | ···················· | ···················· | ···················· | ···················· |
| Q | 1755 hCoV-19/USA/NY-NYCPHL-000184/2020\|EPI_ISL_631719\|2 | ···················· | ···················· | ···················· | ···················· | ···················· | ···················· |
| Q | 1756 hCoV-19/USA/NY-NYCPHL-000185/2020\|EPI_ISL_631720\|2 | ···················· | ···················· | ···················· | ···················· | ···················· | ···················· |
| Q | 1757 hCoV-19/USA/NY-NYCPHL-000210/2020\|EPI_ISL_631743\|2 | ···················· | ···················· | ···················· | ···················· | ···················· | ···················· |
| Q | 1758 hCoV-19/USA/NY-NYCPHL-000211/2020\|EPI_ISL_631744\|2 | ···················· | ···················· | ···················· | ···················· | ···················· | ···················· |
| Q | 1759 hCoV-19/USA/NY-NYCPHL-000212/2020\|EPI_ISL_631745\|2 | ···················· | ···················· | ···················· | ···················· | ···················· | ···················· |
| Q | 1760 hCoV-19/USA/NY-NYCPHL-000214/2020\|EPI_ISL_631746\|2 | ···················· | ···················· | ···················· | ···················· | ···················· | ···················· |
| Q | 1761 hCoV-19/USA/NY-NYCPHL-000216/2020\|EPI_ISL_631747\|2 | ···················· | ···················· | ···················· | ···················· | ···················· | ···················· |
| Q | 1762 hCoV-19/USA/NY-NYCPHL-000218/2020\|EPI_ISL_631749\|2 | ···················· | ···················· | ···················· | ···················· | ···················· | ···················· |
| Q | 1763 hCoV-19/USA/NY-NYCPHL-000219/2020\|EPI_ISL_631750\|2 | ···················· | ···················· | ···················· | ···················· | ···················· | ···················· |
| Q | 1764 hCoV-19/USA/NY-NYCPHL-000220/2020\|EPI_ISL_631751\|2 | ···················· | ···················· | ···················· | ···················· | ···················· | ···················· |
| Q | 1765 hCoV-19/USA/NY-NYCPHL-000222/2020\|EPI_ISL_631752\|2 | ···················· | ···················· | ···················· | ···················· | ···················· | ···················· |
| Q | 1766 hCoV-19/USA/NY-NYCPHL-000223/2020\|EPI_ISL_631753\|2 | ···················· | ···················· | ···················· | ···················· | ···················· | ···················· |
| Q | 1767 hCoV-19/USA/NY-NYCPHL-000922/2020\|EPI_ISL_632977\|2 | ···················· | ···················· | ···················· | ···················· | ···················· | ···················· |
| Q | 1768 hCoV-19/USA/NY-NYCPHL-000925/2020\|EPI_ISL_632979\|2 | ···················· | ···················· | ···················· | ···················· | ···················· | ···················· |
| Q | 1769 hCoV-19/USA/NY-NYCPHL-000938/2020\|EPI_ISL_632980\|2 | ···················· | ···················· | ···················· | ···················· | ···················· | ···················· |
| Q | 1770 hCoV-19/USA/NY-NYCPHL-000940/2020\|EPI_ISL_632981\|2 | ···················· | ·A·················· | ···················· | ···················· | ···················· | ···················· |
| Q | 1771 hCoV-19/USA/NY-NYCPHL-000943/2020\|EPI_ISL_632982\|2 | ···················· | ···················· | ···················· | ···················· | ···················· | ···················· |
| Q | 1772 hCoV-19/USA/NY-NYCPHL-000946/2020\|EPI_ISL_632984\|2 | ···················· | ···················· | ···················· | ···················· | ···················· | ···················· |
| Q | 1773 hCoV-19/USA/NY-NYCPHL-000947/2020\|EPI_ISL_632985\|2 | ···················· | ···················· | ···················· | ···················· | ···················· | ···················· |
| Q | 1774 hCoV-19/USA/NY-NYCPHL-000948/2020\|EPI_ISL_632986\|2 | ···················· | ···················· | ···················· | ···················· | ···················· | ···················· |
| Q | 1775 hCoV-19/USA/NY-NYCPHL-000950/2020\|EPI_ISL_632987\|2 | ···················· | ···················· | ···················· | ···················· | ···················· | ···················· |
| Q | 1776 hCoV-19/USA/NY-NYCPHL-000952/2020\|EPI_ISL_632989\|2 | ···················· | ···················· | ···················· | ···················· | ···················· | ···················· |
| Q | 1777 hCoV-19/USA/NY-NYCPHL-000953/2020\|EPI_ISL_632990\|2 | ···················· | ···················· | ···················· | ···················· | ···················· | ···················· |
| Q | 1778 hCoV-19/USA/NY-NYCPHL-000954/2020\|EPI_ISL_632991\|2 | ···················· | ···················· | ···················· | ···················· | ···················· | ···················· |
| Q | 1779 hCoV-19/USA/NY-NYCPHL-000956/2020\|EPI_ISL_632992\|2 | ···················· | ···················· | ···················· | ···················· | ···················· | ···················· |
| Q | 1780 hCoV-19/USA/NY-NYCPHL-000957/2020\|EPI_ISL_632993\|2 | ···················· | ···················· | ···················· | ···················· | ···················· | ···················· |
| Q | 1781 hCoV-19/USA/NY-NYCPHL-000959/2020\|EPI_ISL_632994\|2 | ···················· | ···················· | ···················· | ···················· | ···················· | ···················· |
| Q | 1782 hCoV-19/USA/NY-NYCPHL-000962/2020\|EPI_ISL_632995\|2 | ···················· | ····C··············· | ···················· | ···················· | ···················· | ···················· |
| Q | 1783 hCoV-19/USA/NY-NYCPHL-000963/2020\|EPI_ISL_632996\|2 | ···················· | ·A·················· | ···················· | ···················· | ···················· | ···················· |
| Q | 1784 hCoV-19/Slovenia/V0-440/2020\|EPI_ISL_635290\|2020-0 | ···················· | ···················· | ···················· | ···················· | ···················· | ···················· |
| Q | 1785 hCoV-19/Slovenia/V0-498/2020\|EPI_ISL_635291\|2020-0 | ···················· | ···················· | ···················· | ···················· | ···················· | ···················· |
| Q | 1786 hCoV-19/Slovenia/V0-5244/2020\|EPI_ISL_635292\|2020- | ···················· | ···················· | ···················· | ···················· | ···················· | ···················· |
| Q | 1787 hCoV-19/Slovenia/V0-571/2020\|EPI_ISL_635293\|2020-0 | ···················· | ···················· | ···················· | ···················· | ···················· | ···················· |
| Q | 1788 hCoV-19/Slovenia/V0-6068/2020\|EPI_ISL_635294\|2020- | ···················· | ···················· | ···················· | ···················· | ···················· | ···················· |
| Q | 1790 hCoV-19/Slovenia/V0-6779/2020\|EPI_ISL_635296\|2020- | ···················· | ···················· | ···················· | ···················· | ···················· | ···················· |
| Q | 1791 hCoV-19/Slovenia/V0-6934/2020\|EPI_ISL_635297\|2020- | ···················· | ···················· | ···················· | ···················· | ···················· | ···················· |
| Q | 1792 hCoV-19/Slovenia/V0-9168/2020\|EPI_ISL_635299\|2020- | ···················· | ···················· | ···················· | ···················· | ···················· | ···················· |
| Q | 1793 hCoV-19/Wales/PHWC-4843E9/2020\|EPI_ISL_628000\|2020 | ···················· | ···················· | ···················· | ···················· | ···················· | ···················· |
| Q | 1797 hCoV-19/Wales/PHWC-484431/2020\|EPI_ISL_628004\|2020 | ···················· | ···················· | ···················· | ···················· | ···················· | ···················· |
| Q | 1798 hCoV-19/Wales/PHWC-48446E/2020\|EPI_ISL_628005\|2020 | ···················· | ···················· | ···················· | ···················· | ···················· | ···················· |
| Q | 1799 hCoV-19/Wales/PHWC-4845C5/2020\|EPI_ISL_628020\|2020 | ···················· | ···················· | ···················· | ···················· | ···················· | ···················· |
| Q | 1800 hCoV-19/Wales/PHWC-4845D4/2020\|EPI_ISL_628021\|2020 | ···················· | ···················· | ···················· | ···················· | ···················· | ···················· |
| Q | 1801 hCoV-19/Wales/PHWC-4845F2/2020\|EPI_ISL_628022\|2020 | ···················· | ···················· | ···················· | ···················· | ···················· | ···················· |
| Q | 1802 hCoV-19/Wales/PHWC-48461D/2020\|EPI_ISL_628023\|2020 | ···················· | ···················· | ···················· | ···················· | ······T············· | ···················· |
| Q | 1803 hCoV-19/Wales/PHWC-48462C/2020\|EPI_ISL_628024\|2020 | ···················· | ···················· | ···················· | ···················· | ···················· | ···················· |
| Q | 1804 hCoV-19/Wales/PHWC-48463B/2020\|EPI_ISL_628025\|2020 | ···················· | ···················· | ···················· | ···················· | ···················· | ···················· |
| Q | 1806 hCoV-19/Wales/PHWC-484659/2020\|EPI_ISL_628027\|2020 | ···················· | ···················· | ···················· | ···················· | ···················· | ···················· |
| Q | 1807 hCoV-19/Wales/PHWC-484668/2020\|EPI_ISL_628028\|2020 | ···················· | ···················· | ···················· | ···················· | ···················· | ···················· |
| Q | 1808 hCoV-19/Wales/PHWC-484677/2020\|EPI_ISL_628029\|2020 | ···················· | ···················· | ···················· | ···················· | ···················· | ···················· |
| Q | 1809 hCoV-19/Wales/PHWC-484686/2020\|EPI_ISL_628030\|2020 | ···················· | ···················· | ···················· | ···················· | ···················· | ···················· |
| Q | 1810 hCoV-19/Wales/PHWC-484695/2020\|EPI_ISL_628031\|2020 | ···················· | ···················· | ···················· | ···················· | ···················· | ···················· |
| Q | 1811 hCoV-19/Wales/PHWC-4846A4/2020\|EPI_ISL_628032\|2020 | ···················· | ···················· | ···················· | ···················· | ···················· | ···················· |
| Q | 1812 hCoV-19/Wales/PHWC-4846C2/2020\|EPI_ISL_628033\|2020 | ···················· | ···················· | ···················· | ···················· | ···················· | ···················· |
| Q | 1814 hCoV-19/Wales/PHWC-4846FF/2020\|EPI_ISL_628035\|2020 | ···················· | ···················· | ···················· | ···················· | ···················· | ···················· |
| Q | 1815 hCoV-19/Wales/PHWC-48471A/2020\|EPI_ISL_628036\|2020 | ···················· | ···················· | ···················· | ···················· | ···················· | ···················· |
| Q | 1816 hCoV-19/Wales/PHWC-484729/2020\|EPI_ISL_628037\|2020 | ···················· | ···················· | ···················· | ···················· | ···················· | ···················· |
| Q | 1817 hCoV-19/Wales/PHWC-484738/2020\|EPI_ISL_628038\|2020 | ···················· | ···················· | ···················· | ···················· | ···················· | ···················· |
| Q | 1818 hCoV-19/Wales/PHWC-484747/2020\|EPI_ISL_628039\|2020 | ···················· | ···················· | ···················· | ···················· | ···················· | ···················· |
| Q | 1819 hCoV-19/Wales/PHWC-484B1E/2020\|EPI_ISL_628080\|2020 | ···················· | ···················· | ···················· | ···················· | ···················· | ···················· |
| Q | 1820 hCoV-19/Wales/PHWC-484B4B/2020\|EPI_ISL_628081\|2020 | ···················· | ···················· | ···················· | ···················· | ···················· | ···················· |
| Q | 1821 hCoV-19/Wales/PHWC-484B5A/2020\|EPI_ISL_628082\|2020 | ···················· | ···················· | ···················· | ···················· | ···················· | ···················· |
| Q | 1822 hCoV-19/Wales/PHWC-484B69/2020\|EPI_ISL_628083\|2020 | ···················· | ···················· | ···················· | ···················· | ···················· | ···················· |
| Q | 1823 hCoV-19/Wales/PHWC-484B87/2020\|EPI_ISL_628084\|2020 | ···················· | ···················· | ···················· | ···················· | ···················· | ···················· |
| Q | 1824 hCoV-19/Wales/PHWC-484BA5/2020\|EPI_ISL_628085\|2020 | ···················· | ···················· | ···················· | ···················· | ···················· | ···················· |
| Q | 1825 hCoV-19/Wales/PHWC-484BB4/2020\|EPI_ISL_628086\|2020 | ···················· | ···················· | ···················· | ···················· | ···················· | ···················· |
| Q | 1826 hCoV-19/Wales/PHWC-484BC3/2020\|EPI_ISL_628087\|2020 | ···················· | ···················· | ···················· | ···················· | ···················· | ···················· |
| Q | 1828 hCoV-19/Wales/PHWC-484C2A/2020\|EPI_ISL_628089\|2020 | ···················· | ···················· | ···················· | ···················· | ···················· | ···················· |
| Q | 1829 hCoV-19/Wales/PHWC-484C39/2020\|EPI_ISL_628090\|2020 | ···················· | ···················· | ···················· | ···················· | ···················· | ···················· |
| Q | 1830 hCoV-19/Wales/PHWC-484C48/2020\|EPI_ISL_628091\|2020 | ···················· | ···················· | ···················· | ···················· | ···················· | ···················· |
| Q | 1832 hCoV-19/Wales/PHWC-484C66/2020\|EPI_ISL_628093\|2020 | ···················· | ···················· | ···················· | ···················· | ···················· | ···················· |
| Q | 1833 hCoV-19/Wales/PHWC-484C84/2020\|EPI_ISL_628094\|2020 | ···················· | ···················· | ···················· | ···················· | ···················· | ···················· |
| Q | 1834 hCoV-19/Wales/PHWC-484CA2/2020\|EPI_ISL_628095\|2020 | ···················· | ···················· | ···················· | ···················· | ···················· | ···················· |
| Q | 1835 hCoV-19/Wales/PHWC-484CB1/2020\|EPI_ISL_628096\|2020 | ···················· | ···················· | ···················· | ···················· | ···················· | ···················· |
| Q | 1837 hCoV-19/Wales/PHWC-484CEE/2020\|EPI_ISL_628098\|2020 | ···················· | ···················· | ···················· | ···················· | ···················· | ···················· |
| Q | 1838 hCoV-19/Wales/PHWC-484CFD/2020\|EPI_ISL_628099\|2020 | ···················· | ···················· | ···················· | ···················· | ···················· | ···················· |
| Q | 1839 hCoV-19/England/MILK-ACEC4C/2020\|EPI_ISL_629330\|20 | ···················· | ···················· | ···················· | ···················· | ···················· | ···················· |
| Q | 1840 hCoV-19/England/MILK-ACC367/2020\|EPI_ISL_629332\|20 | ···················· | ···················· | ···················· | ···················· | ···················· | ···················· |
| Q | 1841 hCoV-19/England/MILK-ACC8B3/2020\|EPI_ISL_629333\|20 | ···················· | ···················· | ···················· | ···················· | ···················· | ···················· |

Q 1842 hCoV-19/England/MILK-AC843B/2020|EPI_ISL_629334|20 ···················· ·A·················· ····················· ···················· ···················· ··A·················
Q 1843 hCoV-19/England/MILK-ACE963/2020|EPI_ISL_629335|20 ···················· ···················· ····················· ···················· ···················· ····················
Q 1844 hCoV-19/England/MILK-ACC798/2020|EPI_ISL_629336|20 ···················· ···················· ····················· ···················· ···················· ····················
Q 1845 hCoV-19/England/MILK-ACBC6D/2020|EPI_ISL_629350|20 ···················· ···················· ····················· ···················· ···················· ····················
Q 1846 hCoV-19/England/MILK-AC95FB/2020|EPI_ISL_629351|20 ···················· ···················· ····················· ···················· ···················· ····················
Q 1847 hCoV-19/England/MILK-ACC15E/2020|EPI_ISL_629352|20 ···················· ···················· ····················· ···················· ·····T·············· ····················
Q 1848 hCoV-19/England/MILK-ACC77A/2020|EPI_ISL_629353|20 ···················· ···················· ····················· ···················· ···················· ····················
Q 1849 hCoV-19/England/MILK-ACE453/2020|EPI_ISL_629354|20 ···················· ···················· ····················· ···················· ···················· ····················
Q 1850 hCoV-19/England/MILK-ACBD97/2020|EPI_ISL_629355|20 ···················· ···················· ····················· ···················· ···················· ····················
Q 1851 hCoV-19/England/MILK-ACBE3A/2020|EPI_ISL_629356|20 ···················· ···················· ····················· ···················· ···················· ····················
Q 1852 hCoV-19/England/MILK-ACEBC7/2020|EPI_ISL_629357|20 ···················· ···················· ····················· ···················· ·····T·············· ····················
Q 1853 hCoV-19/England/MILK-AC7D31/2020|EPI_ISL_629358|20 ···················· ···················· ····················· ···················· ···················· ····················
Q 1854 hCoV-19/England/MILK-ACE8DF/2020|EPI_ISL_629359|20 ···················· ···················· ····················· ···················· ···················· ····················
Q 1855 hCoV-19/England/MILK-ACE936/2020|EPI_ISL_629360|20 ···················· ···················· ····················· ···················· ···················· ····················
Q 1856 hCoV-19/England/MILK-ACE3A1/2020|EPI_ISL_629361|20 ···················· ···················· ····················· ···················· ···················· ····················
Q 1857 hCoV-19/England/MILK-ACBB60/2020|EPI_ISL_629362|20 ···················· ···················· ····················· ···················· ···················· ····················
Q 1858 hCoV-19/England/MILK-AC85ED/2020|EPI_ISL_629364|20 ···················· ···················· ····················· ···················· ···················· ····················
Q 1859 hCoV-19/England/MILK-ACC9A1/2020|EPI_ISL_629365|20 ···················· ···················· ····················· ···················· ···················· ····················
Q 1860 hCoV-19/England/MILK-ACE4F9/2020|EPI_ISL_629366|20 ···················· ···················· ····················· ···················· ···················· ····················
Q 1861 hCoV-19/England/MILK-AC8495/2020|EPI_ISL_629367|20 ···················· ···················· ····················· ···················· ···················· ····················
Q 1862 hCoV-19/England/MILK-ACC9B0/2020|EPI_ISL_629368|20 ···················· ···················· ····················· ···················· ···················· ····················
Q 1863 hCoV-19/England/MILK-ACE21D/2020|EPI_ISL_629369|20 ···················· ···················· ····················· ···················· ···················· ····················
Q 1864 hCoV-19/England/MILK-ACDFBD/2020|EPI_ISL_630313|20 ···················· ···················· ····················· ···················· ···················· ····················
Q 1865 hCoV-19/England/MILK-ACCD87/2020|EPI_ISL_630314|20 ···················· ·A·················· ····················· ···················· ···················· ··A·················
Q 1866 hCoV-19/England/MILK-ACDD77/2020|EPI_ISL_630315|20 ···················· ···················· ····················· ···················· ···················· ····················
Q 1867 hCoV-19/England/MILK-AC73B7/2020|EPI_ISL_630316|20 ···················· ···················· ····················· ···················· ···················· ····················
Q 1868 hCoV-19/England/MILK-ACCD5A/2020|EPI_ISL_630335|20 ···················· ···················· ····················· ···················· ···················· ····················
Q 1869 hCoV-19/England/MILK-ACD07E/2020|EPI_ISL_630336|20 ···················· ···················· ····················· ···················· ···················· ····················
Q 1870 hCoV-19/England/MILK-ACCFDC/2020|EPI_ISL_630337|20 ···················· ···················· ····················· ···················· ···················· ····················
Q 1871 hCoV-19/England/MILK-ACCC3F/2020|EPI_ISL_630338|20 ···················· ···················· ····················· ···················· ···················· ····················
Q 1872 hCoV-19/England/MILK-ACD14E/2020|EPI_ISL_630339|20 ···················· ·A·················· ····················· ···················· ···················· ··A·················
Q 1873 hCoV-19/England/MILK-ACDAF8/2020|EPI_ISL_630340|20 ···················· ···················· ····················· ···················· ···················· ····················
Q 1874 hCoV-19/England/MILK-ACD0D8/2020|EPI_ISL_630341|20 ···················· ···················· ····················· ···················· ···················· ····················
Q 1875 hCoV-19/England/MILK-AC7320/2020|EPI_ISL_630342|20 ···················· ·A······A··········· ····················· ···················· ···················· ····················
Q 1876 hCoV-19/England/MILK-ACDD59/2020|EPI_ISL_630344|20 ···················· ···················· ····················· ···················· ···················· ····················
Q 1877 hCoV-19/England/MILK-AC734E/2020|EPI_ISL_630346|20 ···················· ···················· ····················· ···················· ···················· ····················
Q 1878 hCoV-19/England/MILK-ACCC4E/2020|EPI_ISL_630347|20 ···················· ···················· ····················· ···················· ···················· ····················
Q 1879 hCoV-19/England/ALDP-A66C91/2020|EPI_ISL_630348|20 ···················· ···················· ····················· ···················· ···················· ····················
Q 1880 hCoV-19/England/ALDP-A66FC5/2020|EPI_ISL_630349|20 ···················· ···················· ····················· ···················· ···················· ····················
Q 1881 hCoV-19/England/ALDP-A671A0/2020|EPI_ISL_630350|20 ···················· ···················· ····················· ···················· ···················· ····················
Q 1882 hCoV-19/England/ALDP-A676DE/2020|EPI_ISL_630351|20 ···················· ···················· ····················· ···················· ···················· ····················
Q 1883 hCoV-19/England/ALDP-A686EC/2020|EPI_ISL_630352|20 ···················· ···················· ····················· ···················· ···················· ····················
Q 1884 hCoV-19/England/ALDP-A67B39/2020|EPI_ISL_630353|20 ···················· ···················· ····················· ···················· ···················· ····················
Q 1885 hCoV-19/England/ALDP-A676A1/2020|EPI_ISL_630354|20 ···················· ·A·················· ····················· ···················· ···················· ····················
Q 1886 hCoV-19/England/ALDP-A677EA/2020|EPI_ISL_630355|20 ···················· ···················· ····················· ···················· ···················· ····················
Q 1887 hCoV-19/England/ALDP-A67559/2020|EPI_ISL_630356|20 ···················· ···················· ····················· ···················· ···················· ····················
Q 1888 hCoV-19/USA/TX-DSHS-1208/2020|EPI_ISL_631669|2020- ···················· ···················· ····················· ···················· ···················· ····················
Q 1889 hCoV-19/USA/TX-DSHS-1223/2020|EPI_ISL_631683|2020- ···················· ···················· ····················· ···················· ···················· ····················
Q 1890 hCoV-19/USA/NY-NYCPHL-000149/2020|EPI_ISL_631685|2 ···················· ···················· ····················· ···················· ···················· ····················
Q 1891 hCoV-19/USA/NY-NYCPHL-000150/2020|EPI_ISL_631686|2 ···················· ···················· ····················· ···················· ···················· ····················
Q 1892 hCoV-19/USA/NY-NYCPHL-000151/2020|EPI_ISL_631687|2 ···················· ···················· ····················· ···················· ···················· ····················
Q 1893 hCoV-19/USA/NY-NYCPHL-000921/2020|EPI_ISL_632976|2 ···················· ···················· ····················· ···················· ···················· ····················
Q 1894 hCoV-19/USA/NY-NYCPHL-000964/2020|EPI_ISL_632997|2 ···················· ···················· ····················· ···················· ···················· ····················
Q 1895 hCoV-19/USA/NY-NYCPHL-000966/2020|EPI_ISL_632998|2 ···················· ···················· ····················· ···················· ···················· ····················
Q 1896 hCoV-19/USA/NY-NYCPHL-000968/2020|EPI_ISL_632999|2 ···················· ···················· ····················· ···················· ···················· ····················
Q 1897 hCoV-19/Norway/4588/2020|EPI_ISL_635100|2020-10-18 ···················· ···················· ····················· ···················· ···················· ····················
Q 1898 hCoV-19/Norway/4619/2020|EPI_ISL_635101|2020-10-18 ···················· ···················· ····················· ···················· ···················· ····················
Q 1899 hCoV-19/Norway/4635/2020|EPI_ISL_635102|2020-10-19 ···················· ···················· ····················· ···················· ···················· ····················
Q 1900 hCoV-19/Norway/4549/2020|EPI_ISL_635103|2020-10-18 ···················· ···················· ····················· ···················· ···················· ····················
Q 1901 hCoV-19/Norway/3364/2020|EPI_ISL_635104|2020-09-03 ···················· ···················· ····················· ···················· ···················· ····················
Q 1902 hCoV-19/Norway/3383/2020|EPI_ISL_635105|2020-08-18 ···················· ·A·················· ····················· ···················· ···················· ··A·················
Q 1903 hCoV-19/Norway/3384/2020|EPI_ISL_635106|2020-08-18 ···················· ···················· ····················· ···················· ···················· ····················
Q 1904 hCoV-19/Norway/3720/2020|EPI_ISL_635107|2020-08-28 ···················· ···················· ····················· ···················· ···················· ····················
Q 1905 hCoV-19/Norway/3723/2020|EPI_ISL_635108|2020-09-09 ···················· ···················· ····················· ···················· ···················· ····················
Q 1906 hCoV-19/Norway/4034/2020|EPI_ISL_635120|2020-09-29 ···················· ···················· ····················· ···················· ···················· ····················
Q 1907 hCoV-19/Norway/3916/2020|EPI_ISL_635121|2020-09-28 ···················· ···················· ····················· ···················· ···················· ····················
Q 1908 hCoV-19/Norway/3919/2020|EPI_ISL_635122|2020-09-29 ···················· ···················· ····················· ···················· ···················· ····················
Q 1909 hCoV-19/Norway/4042/2020|EPI_ISL_635130|2020-09-24 ···················· ···················· ····················· ···················· ···················· ····················
Q 1910 hCoV-19/Norway/4045/2020|EPI_ISL_635131|2020-09-24 ···················· ···················· ····················· ···················· ···················· ····················
Q 1911 hCoV-19/Norway/4067/2020|EPI_ISL_635132|2020-09-25 ···················· ···A················ ····················· ···················· ···················· ····················
Q 1912 hCoV-19/Norway/4109/2020|EPI_ISL_635133|2020-09-21 ···················· ···················· ····················· ···················· ···················· ····················
Q 1913 hCoV-19/Norway/4065/2020|EPI_ISL_635134|2020-10-02 ···················· ···················· ····················· ···················· ···················· ····················
Q 1914 hCoV-19/Norway/4058/2020|EPI_ISL_635135|2020-09-30 ···················· ···················· ····················· ···················· ···················· ····················
Q 1915 hCoV-19/Norway/4059/2020|EPI_ISL_635136|2020-09-30 ···················· ···················· ······A·············· ···················· ···················· ····················
Q 1916 hCoV-19/Norway/4098/2020|EPI_ISL_635137|2020-10-01 ···················· ···················· ····················· ···················· ···················· ····················
Q 1917 hCoV-19/Norway/4049/2020|EPI_ISL_635138|2020-10-02 ···················· ···················· ····················· ···················· ···················· ····················
Q 1918 hCoV-19/Norway/4343/2020|EPI_ISL_635139|2020-10-07 ···················· ···················· ····················· ···················· ···················· ····················
Q 1919 hCoV-19/Norway/4347/2020|EPI_ISL_635140|2020-10-10 ···················· ···················· ····················· ···················· ·····T·············· ····················
Q 1920 hCoV-19/Norway/4339/2020|EPI_ISL_635141|2020-10-12 ···················· ···················· ····················· ···················· ···················· ····················
Q 1921 hCoV-19/Norway/4337/2020|EPI_ISL_635142|2020-10-12 ···················· ···················· ····················· ···················· ···················· ····················

```
Q 1922 hCoV-19/Norway/4276/2020|EPI_ISL_635143|2020-10-10 ···················· ···················· ····················· ···················· ···················· ····················
Q 1923 hCoV-19/Norway/4236/2020|EPI_ISL_635144|2020-10-06 ···················· ···················· ····················· ···················· ···················· ····················
Q 1924 hCoV-19/Norway/4333/2020|EPI_ISL_635145|2020-10-12 ···················· ···················· ····················· ···················· ·····T·············· ····················
Q 1925 hCoV-19/Norway/3808/2020|EPI_ISL_635146|2020-08-10 ···················· ·A·················· ····················· ···················· ···················· ··A·················
Q 1926 hCoV-19/Norway/3599/2020|EPI_ISL_635147|2020-07-31 ···················· ·A·················· ····················· ···················· ···················· ··A·················
Q 1927 hCoV-19/Norway/3565/2020|EPI_ISL_635148|2020-08-01 ···················· ·A·················· ····················· ···················· ···················· ··A·················
Q 1928 hCoV-19/Norway/3783/2020|EPI_ISL_635149|2020-08-31 ···················· ···················· ····················· ···················· ···················· ····················
Q 1929 hCoV-19/Norway/3769/2020|EPI_ISL_635150|2020-09-02 ···················· ···················· ····················· ···················· ···················· ····················
Q 1930 hCoV-19/Norway/3773/2020|EPI_ISL_635151|2020-09-04 ···················· ···················· ····················· ···················· ···················· ····················
Q 1931 hCoV-19/Norway/3776/2020|EPI_ISL_635152|2020-09-08 ···················· ···················· ····················· ···················· ···················· ····················
Q 1932 hCoV-19/Norway/4328/2020|EPI_ISL_635153|2020-10-07 ···················· ···················· ····················· ···················· ···················· ····················
Q 1933 hCoV-19/Norway/4329/2020|EPI_ISL_635154|2020-10-05 ···················· ···················· ····················· ···················· ···················· ····················
Q 1934 hCoV-19/Norway/4122/2020|EPI_ISL_635155|2020-09-30 ···················· ···················· ····················· ···················· ···················· ····················
Q 1935 hCoV-19/Norway/3579/2020|EPI_ISL_635156|2020-07-31 ···················· ·A·················· ····················· ···················· ···················· ··A·················
Q 1936 hCoV-19/Norway/3586/2020|EPI_ISL_635157|2020-07-31 ···················· ·A·················· ····················· ···················· ···················· ··A·················
Q 1937 hCoV-19/Norway/4069/2020|EPI_ISL_635158|2020-09-28 ···················· ···················· ····················· ···················· ···················· ····················
Q 1938 hCoV-19/Norway/4066/2020|EPI_ISL_635159|2020-10-04 ···················· ···················· ····················· ···················· ···················· ····················
Q 1939 hCoV-19/Norway/4342/2020|EPI_ISL_635160|2020-10-12 ···················· ···················· ····················· ···················· ···················· ····················
Q 1940 hCoV-19/Norway/4640/2020|EPI_ISL_635161|2020-10-18 ···················· ···················· ····················· ···················· ···················· ····················
Q 1941 hCoV-19/Norway/4596/2020|EPI_ISL_635162|2020-10-12 ···················· ···················· ····················· ···················· ·····T·············· ····················
Q 1942 hCoV-19/Norway/4332/2020|EPI_ISL_635163|2020-10-12 ···················· ···················· ····················· ···················· ···················· ····················
Q 1943 hCoV-19/Norway/4631/2020|EPI_ISL_635164|2020-10-18 ···················· ···················· ····················· ···················· ···················· ····················
Q 1944 hCoV-19/Norway/4590/2020|EPI_ISL_635165|2020-10-18 ···················· ···················· ····················· ···················· ···················· ····················
Q 1945 hCoV-19/Norway/4235/2020|EPI_ISL_635166|2020-10-09 ···················· ···················· ····················· ···················· ···················· ····················
Q 1946 hCoV-19/Norway/4627/2020|EPI_ISL_635167|2020-10-12 ···················· ···················· ····················· ···················· ···················· ····················
Q 1947 hCoV-19/Norway/4543/2020|EPI_ISL_635168|2020-10-15 ···················· ···················· ····················· ···················· ···················· ····················
Q 1948 hCoV-19/Norway/4622/2020|EPI_ISL_635169|2020-10-18 ···················· ···················· ····················· ···················· ···················· ····················
Q 1949 hCoV-19/Slovenia/810/2020|EPI_ISL_635201|2020-03-0 ···················· ···················· ····················· ···················· ···················· ····················
Q 1950 hCoV-19/Slovenia/4384/2020|EPI_ISL_635206|2020-03- ···················· ···················· ····················· ···················· ···················· ····················
Q 1951 hCoV-19/Slovenia/4499/2020|EPI_ISL_635207|2020-03- ···················· ···················· ····················· ···················· ···················· ····················
Q 1952 hCoV-19/Jordan/AM-ALSR-4058/2020|EPI_ISL_636418|20 ···················· ···················· ····················· ···················· ···················· ····················
Q 1953 hCoV-19/Jordan/AM-ALSR-4059/2020|EPI_ISL_636419|20 ···················· ···················· ····················· ···················· ·····T·············· ····················
Q 1954 hCoV-19/Jordan/AM-ALSR-4060/2020|EPI_ISL_636420|20 ···················· ···················· ····················· ···················· ···················· ····················
Q 1955 hCoV-19/Jordan/AM-ALSR-4061/2020|EPI_ISL_636421|20 ···················· ···················· ····················· ···················· ···················· ····················
Q 1956 hCoV-19/Jordan/AM-ALSR-4062/2020|EPI_ISL_636422|20 ···················· ···················· ····················· ···················· ···················· ····················
Q 1957 hCoV-19/Jordan/AM-ALSR-4063/2020|EPI_ISL_636423|20 ···················· ···················· ····················· ···················· ···················· ····················
Q 1958 hCoV-19/Jordan/AM-ALSR-4064/2020|EPI_ISL_636424|20 ···················· ···················· ····················· ···················· ···················· ····················
Q 1959 hCoV-19/Jordan/AM-ALSR-4065/2020|EPI_ISL_636425|20 ···················· ···················· ····················· ···················· ···················· ····················
Q 1960 hCoV-19/Jordan/AM-ALSR-4066/2020|EPI_ISL_636426|20 ···················· ···················· ····················· ···················· ···················· ····················
Q 1961 hCoV-19/Jordan/AM-ALSR-4067/2020|EPI_ISL_636427|20 ···················· ···················· ····················· ···················· ···················· ····················
Q 1962 hCoV-19/Jordan/AM-ALSR-4068/2020|EPI_ISL_636428|20 ···················· ···················· ····················· ···················· ···················· ····················
Q 1963 hCoV-19/Jordan/AM-ALSR-4069/2020|EPI_ISL_636429|20 ···················· ···················· ····················· ···················· ···················· ····················
Q 1964 hCoV-19/Jordan/AM-ALSR-4070/2020|EPI_ISL_636430|20 ···················· ···················· ····················· ···················· ···················· ····················
Q 1965 hCoV-19/Jordan/AM-ALSR-4071/2020|EPI_ISL_636431|20 ···················· ···················· ····················· ···················· ···················· ····················
Q 1966 hCoV-19/Jordan/AM-ALSR-4072/2020|EPI_ISL_636432|20 ···················· ···················· ····················· ···················· ···················· ····················
Q 1967 hCoV-19/Jordan/AM-ALSR-4073/2020|EPI_ISL_636433|20 ···················· ···················· ····················· ···················· ···················· ····················
Q 1968 hCoV-19/Jordan/AM-ALSR-4074/2020|EPI_ISL_636434|20 ···················· ···················· ····················· ···················· ···················· ····················
Q 1969 hCoV-19/Jordan/AM-ALSR-4075/2020|EPI_ISL_636435|20 ···················· ···················· ····················· ···················· ···················· ····················
Q 1970 hCoV-19/Jordan/AM-ALSR-4076/2020|EPI_ISL_636436|20 ···················· ···················· ····················· ···················· ···················· ····················
Q 1971 hCoV-19/Jordan/AM-ALSR-4077/2020|EPI_ISL_636437|20 ···················· ···················· ····················· ···················· ···················· ····················
Q 1972 hCoV-19/Jordan/AM-ALSR-4078/2020|EPI_ISL_636438|20 ···················· ·A·················· ····················· ···················· ···················· ··A·················
Q 1973 hCoV-19/Jordan/AM-ALSR-4079/2020|EPI_ISL_636439|20 ···················· ···················· ····················· ···················· ···················· ····················
Q 1974 hCoV-19/Jordan/AM-ALSR-4094/2020|EPI_ISL_636454|20 ···················· ···················· ····················· ···················· ···················· ····················
Q 1975 hCoV-19/Jordan/AM-ALSR-4095/2020|EPI_ISL_636455|20 ···················· ···················· ····················· ···················· ···················· ····················
Q 1976 hCoV-19/Italy/VEN-IZSVe-10599/2020|EPI_ISL_636462| ···················· ···················· ····················· ···················· ···················· ····················
Q 1977 hCoV-19/Italy/VEN-IZSVe-10860/2020|EPI_ISL_636463| ···················· ···················· ····················· ···················· ···················· ····················
Q 1978 hCoV-19/Italy/VEN-IZSVe-1-1/2020|EPI_ISL_636464|20 ···················· ···················· ····················· ···················· ···················· ····················
Q 1979 hCoV-19/Italy/VEN-IZSVe-14-21/2020|EPI_ISL_636467| ···················· ···················· ····················· ···················· ···················· ····················
Q 1980 hCoV-19/Italy/VEN-IZSVe-14-30/2020|EPI_ISL_636468| ···················· ···················· ····················· ···················· ···················· ····················
Q 1981 hCoV-19/Italy/VEN-IZSVe-14-36/2020|EPI_ISL_636469| ···················· ···················· ····················· ···················· ···················· ····················
Q 1982 hCoV-19/Italy/VEN-IZSVe-14-48/2020|EPI_ISL_636470| ···················· ···················· ····················· ···················· ···················· ····················
Q 1983 hCoV-19/Italy/VEN-IZSVe-20-25/2020|EPI_ISL_636471| ···················· ···················· ····················· ···················· ···················· ····················
Q 1984 hCoV-19/Italy/VEN-IZSVe-2-2/2020|EPI_ISL_636472|20 ···················· ···················· ····················· ···················· ···················· ····················
Q 1985 hCoV-19/Italy/VEN-IZSVe-37-4/2020|EPI_ISL_636473|2 ···················· ···················· ····················· ···················· ···················· ····················
Q 1986 hCoV-19/Italy/VEN-IZSVe-6-47/2020|EPI_ISL_636474|2 ···················· ···················· ····················· ···················· ···················· ····················
Q 1987 hCoV-19/Italy/VEN-IZSVe-6-55/2020|EPI_ISL_636475|2 ···················· ···················· ····················· ···················· ···················· ····················
Q 1988 hCoV-19/France/ARA-32967/2020|EPI_ISL_636476|2020- ···················· ···················· ····················· ···················· ···················· ····················
Q 1989 hCoV-19/France/ARA-33082/2020|EPI_ISL_636477|2020- ···················· ···················· ····················· ···················· ···················· ····················
Q 1990 hCoV-19/France/ARA-33485/2020|EPI_ISL_636478|2020- ···················· ···················· ····················· ···················· ···················· ····················
Q 1991 hCoV-19/France/ARA-33853/2020|EPI_ISL_636479|2020- ···················· ···················A ····················· ···················· ···················· ···················· 1
Q 1992 hCoV-19/France/ARA-34732/2020|EPI_ISL_636480|2020- ···················· ···················· ····················· ···················· ···················· ····················
Q 1993 hCoV-19/France/ARA-34744/2020|EPI_ISL_636481|2020- ···················· ···················· ····················· ···················· ···················· ····················
Q 1994 hCoV-19/France/ARA-36389/2020|EPI_ISL_636482|2020- ···················· ···················· ····················· ···················· ···················· ····················
Q 1995 hCoV-19/France/ARA-36390/2020|EPI_ISL_636483|2020- ···················· ···················· ····················· ···················· ···················· ····················
Q 1996 hCoV-19/France/ARA-36541/2020|EPI_ISL_636484|2020- ···················· ···················· ····················· ···················· ···················· ····················
Q 1997 hCoV-19/France/ARA-36991/2020|EPI_ISL_636485|2020- ···················· ···················· ····················· ···················· ···················· ····················
Q 1998 hCoV-19/France/ARA-37154/2020|EPI_ISL_636486|2020- ···················· ···················· ····················· ···················· ···················· ····················
Q 1999 hCoV-19/France/ARA-33428/2020|EPI_ISL_636487|2020- ···················· ···················· ····················· ···················· ···················· ····················
Q 2000 hCoV-19/Italy/VEN-IZSVe-53842/2020|EPI_ISL_636488| ···················· ···················· ····················· ···················· ···················· ····················
Q 2001 hCoV-19/Netherlands/NH-RIVM-10393/2020|EPI_ISL_636 ···················· ···················· ····················· ···················· ···················· ····················
```

| | | | | | | | |
|---|---|---|---|---|---|---|---|
| Q | 2002 hCoV-19/Curacao/CW-RIVM-10308/2020\|EPI_ISL_636492\| | ···················· | ···················· | ···················· | ···················· | ···················· | ···················· |
| Q | 2003 hCoV-19/Curacao/CW-RIVM-10309/2020\|EPI_ISL_636493\| | ···················· | ···················· | ···················· | ···················· | ···················· | ···················· |
| Q | 2004 hCoV-19/Netherlands/GE-RIVM-10307/2020\|EPI_ISL_636 | ···················· | ···················· | ···················· | ···················· | ···················· | ···················· |
| Q | 2005 hCoV-19/Netherlands/GE-RIVM-10319/2020\|EPI_ISL_636 | ···················· | ···················· | ···················· | ···················· | ···················· | ···················· |
| Q | 2006 hCoV-19/Netherlands/GE-RIVM-10323/2020\|EPI_ISL_636 | ···················· | ···················· | ···················· | ···················· | ···················· | ···················· |
| Q | 2007 hCoV-19/Netherlands/GE-RIVM-10324/2020\|EPI_ISL_636 | ···················· | ···················· | ···················· | ···················· | ···················· | ···················· |
| Q | 2008 hCoV-19/Netherlands/GE-RIVM-10328/2020\|EPI_ISL_636 | ···················· | ···················· | ···················· | ···················· | ···················· | ···················· |
| Q | 2009 hCoV-19/Netherlands/GE-RIVM-10329/2020\|EPI_ISL_636 | ···················· | ···················· | ···················· | ···················· | ···················· | ···················· |
| Q | 2010 hCoV-19/Netherlands/GE-RIVM-10332/2020\|EPI_ISL_636 | ···················· | ···················· | ···················· | ···················· | ···················· | ···················· |
| Q | 2011 hCoV-19/Netherlands/GE-RIVM-10385/2020\|EPI_ISL_636 | ···················· | ···················· | ···················· | ···················· | ···················· | ···················· |
| Q | 2012 hCoV-19/Netherlands/GE-RIVM-10386/2020\|EPI_ISL_636 | ···················· | ···················· | ···················· | ···················· | ···················· | ···················· |
| Q | 2013 hCoV-19/Netherlands/LI-RIVM-10345/2020\|EPI_ISL_636 | ···················· | ···················· | ···················· | ···················· | ···················· | ···················· |
| Q | 2014 hCoV-19/Netherlands/NB-RIVM-10300/2020\|EPI_ISL_636 | ···················· | ···················· | ···················· | ···················· | ···················· | ···················· |
| Q | 2015 hCoV-19/Netherlands/NB-RIVM-10301/2020\|EPI_ISL_636 | ···················· | ···················· | ···················· | ···················· | ···················· | ···················· |
| Q | 2016 hCoV-19/Netherlands/NB-RIVM-10304/2020\|EPI_ISL_636 | ···················· | ···················· | ···················· | ···················· | ···················· | ···················· |
| Q | 2017 hCoV-19/Netherlands/NB-RIVM-10317/2020\|EPI_ISL_636 | ···················· | ···················· | ···················· | ···················· | ···················· | ···················· |
| Q | 2018 hCoV-19/Netherlands/NB-RIVM-10367/2020\|EPI_ISL_636 | ···················· | ···················· | ···················· | ···················· | ···················· | ···················· |
| Q | 2019 hCoV-19/Netherlands/NB-RIVM-10368/2020\|EPI_ISL_636 | ···················· | ···················· | ···················· | ···················· | ···················· | ···················· |
| Q | 2020 hCoV-19/Netherlands/NH-RIVM-10311/2020\|EPI_ISL_636 | ···················· | ···················· | ···················· | ···················· | ···················· | ···················· |
| Q | 2021 hCoV-19/Netherlands/UT-RIVM-10302/2020\|EPI_ISL_636 | ···················· | ···················· | ···················· | ···················· | ···················· | ···················· |
| Q | 2022 hCoV-19/Netherlands/ZH-RIVM-10327/2020\|EPI_ISL_636 | ···················· | ···················· | ···················· | ···················· | ···················· | ···················· |
| Q | 2023 hCoV-19/Aruba/AW-RIVM-10326/2020\|EPI_ISL_636513\|20 | ···················· | ···················· | ···················· | ···················· | ···················· | ···················· |
| Q | 2024 hCoV-19/Aruba/AW-RIVM-10388/2020\|EPI_ISL_636514\|20 | ···················· | ·A·················· | ···················· | ···················· | ···················· | ···················· |
| Q | 2025 hCoV-19/Aruba/AW-RIVM-10389/2020\|EPI_ISL_636515\|20 | ···················· | ·A·················· | ···················· | ···················· | ···················· | ···················· |
| Q | 2026 hCoV-19/Aruba/AW-RIVM-10390/2020\|EPI_ISL_636516\|20 | ···················· | ·A·················· | ···················· | ···················· | ···················· | ···················· |
| Q | 2027 hCoV-19/Aruba/AW-RIVM-10391/2020\|EPI_ISL_636517\|20 | ···················· | ···················· | ···················· | ···················· | ···················· | ···················· |
| Q | 2028 hCoV-19/Aruba/AW-RIVM-10392/2020\|EPI_ISL_636518\|20 | ···················· | ·A·················· | ···················· | ···················· | ···················· | ···················· |
| Q | 2029 hCoV-19/Curacao/CW-RIVM-10364/2020\|EPI_ISL_636519\| | ···················· | ·A·················· | ···················· | ···················· | ···················· | ···················· |
| Q | 2030 hCoV-19/Curacao/CW-RIVM-10365/2020\|EPI_ISL_636520\| | ···················· | ·A·················· | ···················· | ···················· | ···················· | ···················· |
| Q | 2031 hCoV-19/Curacao/CW-RIVM-10366/2020\|EPI_ISL_636521\| | ···················· | ·A·················· | ···················· | ···················· | ···················· | ···················· |
| Q | 2032 hCoV-19/Netherlands/FL-RIVM-10336/2020\|EPI_ISL_636 | ···················· | ···················· | ···················· | ···················· | ···················· | ···················· |
| Q | 2033 hCoV-19/Netherlands/FL-RIVM-10337/2020\|EPI_ISL_636 | ···················· | ···················· | ···················· | ···················· | ···················· | ···················· |
| Q | 2034 hCoV-19/Netherlands/FL-RIVM-10338/2020\|EPI_ISL_636 | ···················· | ···················· | ···················· | ···················· | ···················· | ···················· |
| Q | 2035 hCoV-19/Netherlands/FL-RIVM-10346/2020\|EPI_ISL_636 | ···················· | ···················· | ···················· | ···················· | ···················· | ···················· |
| Q | 2036 hCoV-19/Netherlands/GE-RIVM-10292/2020\|EPI_ISL_636 | ···················· | ···················· | ···················· | ···················· | ···················· | ···················· |
| Q | 2037 hCoV-19/Netherlands/GE-RIVM-10303/2020\|EPI_ISL_636 | ···················· | ···················· | ···················· | ···················· | ···················· | ···················· |
| Q | 2038 hCoV-19/Netherlands/GE-RIVM-10314/2020\|EPI_ISL_636 | ···················· | ···················· | ···················· | ···················· | ···················· | ···················· |
| Q | 2039 hCoV-19/Netherlands/GE-RIVM-10320/2020\|EPI_ISL_636 | ···················· | ···················· | ···················· | ···················· | ···················· | ···················· |
| Q | 2040 hCoV-19/Netherlands/GE-RIVM-10321/2020\|EPI_ISL_636 | ···················· | ···················· | ···················· | ···················· | ···················· | ···················· |
| Q | 2041 hCoV-19/Netherlands/GE-RIVM-10322/2020\|EPI_ISL_636 | ···················· | ···················· | ···················· | ···················· | ···················· | ···················· |
| Q | 2042 hCoV-19/Netherlands/GE-RIVM-10325/2020\|EPI_ISL_636 | ···················· | ···················· | ···················· | ···················· | ···················· | ···················· |
| Q | 2043 hCoV-19/Netherlands/GE-RIVM-10374/2020\|EPI_ISL_636 | ···················· | ···················· | ···················· | ···················· | ···················· | ···················· |
| Q | 2044 hCoV-19/Netherlands/GE-RIVM-10378/2020\|EPI_ISL_636 | ···················· | ···················· | ···················· | ···················· | ···················· | ···················· |
| Q | 2045 hCoV-19/Netherlands/GE-RIVM-10379/2020\|EPI_ISL_636 | ···················· | ···················· | ···················· | ···················· | ···················· | ···················· |
| Q | 2046 hCoV-19/Netherlands/GE-RIVM-10382/2020\|EPI_ISL_636 | ···················· | ···················· | ···················· | ···················· | ······T············· | ···················· |
| Q | 2047 hCoV-19/Netherlands/GE-RIVM-10399/2020\|EPI_ISL_636 | ···················· | ···················· | ···················· | ···················· | ···················· | ···················· |
| Q | 2048 hCoV-19/Netherlands/GE-RIVM-10401/2020\|EPI_ISL_636 | ···················· | ···················· | ···················· | ···················· | ···················· | ···················· |
| Q | 2049 hCoV-19/England/MILK-AC835C/2020\|EPI_ISL_629250\|20 | ···················· | ···················· | ···················· | ···················· | ···················· | ···················· |
| Q | 2050 hCoV-19/England/MILK-ACE514/2020\|EPI_ISL_629251\|20 | ···················· | ···················· | ···················· | ···················· | ···················· | ···················· |
| Q | 2051 hCoV-19/England/MILK-ACE338/2020\|EPI_ISL_629254\|20 | ···················· | ···················· | ···················· | ···················· | ···················· | ···················· |
| Q | 2052 hCoV-19/England/MILK-AC9634/2020\|EPI_ISL_629255\|20 | ···················· | ···················· | ···················· | ···················· | ···················· | ···················· |
| Q | 2053 hCoV-19/England/MILK-ACC7A7/2020\|EPI_ISL_629260\|20 | ···················· | ···················· | ···················· | ···················· | ···················· | ···················· |
| Q | 2054 hCoV-19/England/MILK-ACE23B/2020\|EPI_ISL_629261\|20 | ···················· | ···················· | ···················· | ···················· | ···················· | ···················· |
| Q | 2055 hCoV-19/England/MILK-AC7D40/2020\|EPI_ISL_629262\|20 | ···················· | ·A·················· | ···················· | ···················· | ···················· | ···A················ |
| Q | 2056 hCoV-19/England/MILK-ACC8C2/2020\|EPI_ISL_629263\|20 | ···················· | ···················· | ···················· | ···················· | ···················· | ···················· |
| Q | 2057 hCoV-19/England/MILK-ACBFDD/2020\|EPI_ISL_629264\|20 | ···················· | ···················· | ···················· | ···················· | ···················· | ···················· |
| Q | 2058 hCoV-19/England/MILK-AC9528/2020\|EPI_ISL_629265\|20 | ···················· | ···················· | ···················· | ···················· | ···················· | ···················· |
| Q | 2059 hCoV-19/England/MILK-ACECC4/2020\|EPI_ISL_629266\|20 | ···················· | ···················· | ···················· | ···················· | ···················· | ···················· |
| Q | 2060 hCoV-19/England/MILK-ACBF82/2020\|EPI_ISL_629267\|20 | ···················· | ···················· | ···················· | ···················· | ···················· | ···················· |
| Q | 2061 hCoV-19/England/MILK-ACC112/2020\|EPI_ISL_629268\|20 | ···················· | ···················· | ···················· | ···················· | ···················· | ···················· |
| Q | 2062 hCoV-19/England/MILK-ACC9DE/2020\|EPI_ISL_629269\|20 | ···················· | ···················· | ···················· | ···················· | ···················· | ···················· |
| Q | 2063 hCoV-19/England/MILK-ACC060/2020\|EPI_ISL_629300\|20 | ···················· | ···················· | ···················· | ···················· | ······T············· | ···················· |
| Q | 2064 hCoV-19/England/MILK-ACC631/2020\|EPI_ISL_629301\|20 | ···················· | ···················· | ···················· | ···················· | ···················· | ···················· |
| Q | 2065 hCoV-19/England/MILK-ACBE1C/2020\|EPI_ISL_629302\|20 | ···················· | ···················· | ···················· | ···················· | ···················· | ···················· |
| Q | 2066 hCoV-19/England/MILK-AC9670/2020\|EPI_ISL_629303\|20 | ···················· | ···················· | ···················· | ···················· | ···················· | ···················· |
| Q | 2067 hCoV-19/England/MILK-ACEC6A/2020\|EPI_ISL_629304\|20 | ···················· | ···················· | ···················· | ···················· | ···················· | ···················· |
| Q | 2068 hCoV-19/England/MILK-ACBE2B/2020\|EPI_ISL_629306\|20 | ···················· | ···················· | ···················· | ···················· | ···················· | ···················· |
| Q | 2069 hCoV-19/England/MILK-ACBCD6/2020\|EPI_ISL_629307\|20 | ···················· | ···················· | ···················· | ···················· | ···················· | ···················· |
| Q | 2070 hCoV-19/England/MILK-ACECB5/2020\|EPI_ISL_629308\|20 | ···················· | ···················· | ···················· | ···················· | ···················· | ···················· |
| Q | 2071 hCoV-19/England/MILK-AC8608/2020\|EPI_ISL_629310\|20 | ···················· | ···················· | ···················· | ···················· | ···················· | ···················· |
| Q | 2072 hCoV-19/England/MILK-AC950A/2020\|EPI_ISL_629312\|20 | ···················· | ···················· | ···················· | ···················· | ···················· | ···················· |
| Q | 2073 hCoV-19/England/MILK-AC8389/2020\|EPI_ISL_629313\|20 | ···················· | ···················· | ···················· | ···················· | ····T··············· | ···················· |
| Q | 2074 hCoV-19/England/MILK-AC94EF/2020\|EPI_ISL_629314\|20 | ···················· | ·A·················· | ···················· | ···················· | ···················· | ···A················ |
| Q | 2075 hCoV-19/England/MILK-AC7CDA/2020\|EPI_ISL_629315\|20 | ···················· | ···················· | ···················· | ···················· | ···················· | ···················· |
| Q | 2076 hCoV-19/England/MILK-ACC1C7/2020\|EPI_ISL_629316\|20 | ···················· | ·A·················· | ···················· | ···················· | ···················· | ···A················ |
| Q | 2077 hCoV-19/England/MILK-AC82F5/2020\|EPI_ISL_629317\|20 | ···················· | ···················· | ···················· | ···················· | ······T············· | ···················· |
| Q | 2078 hCoV-19/England/MILK-AC85A1/2020\|EPI_ISL_629318\|20 | ···················· | ···················· | ···················· | ···················· | ···················· | ···················· |
| Q | 2079 hCoV-19/England/MILK-ACCA08/2020\|EPI_ISL_629319\|20 | ···················· | ···················· | ···················· | ···················· | ···················· | ···················· |
| Q | 2080 hCoV-19/England/MILK-AC95DD/2020\|EPI_ISL_629321\|20 | ···················· | ···················· | ···················· | ···················· | ···················· | ···················· |
| Q | 2081 hCoV-19/England/MILK-ACBE67/2020\|EPI_ISL_629322\|20 | ···················· | ···················· | ···················· | ···················· | ···················· | ···················· |

Q 2082 hCoV-19/England/MILK-ACC40A/2020|EPI_ISL_629324|20 .................... .................... ..................... .................... .................... ....................
Q 2083 hCoV-19/England/MILK-ACED0D/2020|EPI_ISL_629325|20 .................... .................... ..................... .................... .................... ....................
Q 2084 hCoV-19/England/MILK-ACC26A/2020|EPI_ISL_629326|20 .................... .A.................. ..................... .................... .................... ..A.................
Q 2085 hCoV-19/England/MILK-AC7D7D/2020|EPI_ISL_629327|20 .................... .................... ..................... .................... .................... ....................
Q 2086 hCoV-19/England/MILK-ACED1C/2020|EPI_ISL_629328|20 .................... .................... ..................... .................... .................... ....................
Q 2087 hCoV-19/England/MILK-ACC033/2020|EPI_ISL_629329|20 .................... .................... ..................... .................... .................... ....................
Q 2088 hCoV-19/England/MILK-ACDA9E/2020|EPI_ISL_630236|20 .................... .................... ..................... .................... .....T.............. ....................
Q 2089 hCoV-19/England/MILK-AC936A/2020|EPI_ISL_630237|20 .................... .................... ..................... .................... .................... ....................
Q 2090 hCoV-19/England/MILK-ACDD86/2020|EPI_ISL_630240|20 .................... .A.................. ..................... .................... .................... ..A.................
Q 2091 hCoV-19/England/MILK-AC736C/2020|EPI_ISL_630241|20 .................... .................... ..................... .................... .................... ....................
Q 2092 hCoV-19/England/MILK-ACCDB4/2020|EPI_ISL_630242|20 .................... .................... ..................... .................... .................... ....................
Q 2093 hCoV-19/England/MILK-AC784F/2020|EPI_ISL_630243|20 .................... .................... ..................... .................... .................... ....................
Q 2094 hCoV-19/England/MILK-ACCD4B/2020|EPI_ISL_630246|20 .................... .................... ..................... .................... .................... ....................
Q 2095 hCoV-19/England/MILK-AC9115/2020|EPI_ISL_630248|20 .................... .................... ..................... .................... .................... ....................
Q 2096 hCoV-19/England/MILK-AC7742/2020|EPI_ISL_630250|20 .................... .................... ..................... .................... .................... ....................
Q 2097 hCoV-19/England/MILK-ACDFCC/2020|EPI_ISL_630251|20 .................... .................... ..................... .................... .................... ....................
Q 2098 hCoV-19/England/MILK-AC72F6/2020|EPI_ISL_630253|20 .................... .................... ..................... .................... .................... ....................
Q 2099 hCoV-19/England/MILK-ACCCE4/2020|EPI_ISL_630256|20 .................... .................... ..................... .................... .................... ....................
Q 2100 hCoV-19/England/MILK-ACD788/2020|EPI_ISL_630257|20 .................... .................... ..................... .................... .................... ....................
Q 2101 hCoV-19/England/MILK-ACD014/2020|EPI_ISL_630300|20 .................... .................... ..................... .................... .................... ....................
Q 2102 hCoV-19/England/MILK-AC7724/2020|EPI_ISL_630302|20 .................... .................... ..................... .................... .................... ....................
Q 2103 hCoV-19/England/MILK-AC74C3/2020|EPI_ISL_630303|20 .................... .................... ..................... .................... .................... ....................
Q 2104 hCoV-19/England/MILK-ACD7E2/2020|EPI_ISL_630304|20 .................... .A.................. ..................... .................... .................... ..A.................
Q 2105 hCoV-19/England/MILK-ACDA61/2020|EPI_ISL_630305|20 .................... .................... ..................... .................... .................... ....................
Q 2106 hCoV-19/England/MILK-ACDFDB/2020|EPI_ISL_630306|20 .................... .................... ..................... .................... .................... ....................
Q 2107 hCoV-19/England/MILK-AC744B/2020|EPI_ISL_630307|20 .................... .................... ..................... .................... .................... ....................
Q 2108 hCoV-19/England/MILK-ACDC89/2020|EPI_ISL_630308|20 .................... .................... ..................... .................... .................... ....................
Q 2109 hCoV-19/England/MILK-AC943A/2020|EPI_ISL_630309|20 .................... .................... ..................... .................... .................... ....................
Q 2110 hCoV-19/England/MILK-ACDF80/2020|EPI_ISL_630310|20 .................... .................... ..................... .................... .....T.............. ....................
Q 2111 hCoV-19/England/MILK-ACDAE9/2020|EPI_ISL_630311|20 .................... .................... ..................... .................... .................... ....................
Q 2112 hCoV-19/England/MILK-ACCDF0/2020|EPI_ISL_630312|20 .................... .................... ..................... .................... .................... ....................
Q 2113 hCoV-19/USA/NY-NYCPHL-000074/2020|EPI_ISL_631567|2 .................... .................... ..................... .................... .................... ....................
Q 2114 hCoV-19/USA/NY-NYCPHL-000075/2020|EPI_ISL_631568|2 .................... .................... ..................... .................... .................... ....................
Q 2115 hCoV-19/USA/NY-NYCPHL-000076/2020|EPI_ISL_631569|2 .................... .................... ..................... .................... .................... ....................
Q 2116 hCoV-19/USA/NY-NYCPHL-000077/2020|EPI_ISL_631570|2 .................... .................... ..................... .................... .................... ....................
Q 2117 hCoV-19/USA/NY-NYCPHL-000078/2020|EPI_ISL_631571|2 .................... .................... ..................... .................... .................... ....................
Q 2118 hCoV-19/USA/NY-NYCPHL-000079/2020|EPI_ISL_631572|2 .................... .................... ..................... .................... .................... ....................
Q 2119 hCoV-19/USA/NY-NYCPHL-000080/2020|EPI_ISL_631573|2 .................... .................... ..................... .................... .................... ....................
Q 2120 hCoV-19/USA/NY-NYCPHL-000081/2020|EPI_ISL_631574|2 .................... .................... ..................... .................... .................... ....................
Q 2121 hCoV-19/USA/NY-NYCPHL-000083/2020|EPI_ISL_631576|2 .................... .................... ..................... .................... .................... ....................
Q 2122 hCoV-19/USA/NY-NYCPHL-000084/2020|EPI_ISL_631577|2 .................... .................... ..................... .................... .................... ....................
Q 2123 hCoV-19/USA/NY-NYCPHL-000085/2020|EPI_ISL_631578|2 .................... .................... ..................... .................... .................... ....................
Q 2124 hCoV-19/USA/NY-NYCPHL-000086/2020|EPI_ISL_631579|2 .................... .................... ..................... .................... .................... ....................
Q 2125 hCoV-19/USA/NY-NYCPHL-000087/2020|EPI_ISL_631580|2 .................... .................... ..................... .................... .................... ....................
Q 2126 hCoV-19/USA/NY-NYCPHL-000088/2020|EPI_ISL_631581|2 .................... ......A............. ..................... .................... .................... ....................
Q 2127 hCoV-19/USA/NY-NYCPHL-000089/2020|EPI_ISL_631582|2 .................... .................... ..................... .................... .................... ....................
Q 2128 hCoV-19/USA/NY-NYCPHL-000090/2020|EPI_ISL_631583|2 .................... .................... ..................... .................... .................... ....................
Q 2129 hCoV-19/USA/NY-NYCPHL-000091/2020|EPI_ISL_631584|2 .................... .................... ..................... .................... .................... ....................
Q 2130 hCoV-19/USA/NY-NYCPHL-000092/2020|EPI_ISL_631585|2 .................... .................... ..................... .................... .................... ....................
Q 2131 hCoV-19/USA/NY-NYCPHL-000093/2020|EPI_ISL_631586|2 .................... .................... ..................... .................... .................... ....................
Q 2132 hCoV-19/USA/NY-NYCPHL-000094/2020|EPI_ISL_631587|2 .................... .................... ..................... .................... .................... ....................
Q 2133 hCoV-19/USA/NY-NYCPHL-000095/2020|EPI_ISL_631588|2 .................... .................... ..................... .................... .................... ....................
Q 2134 hCoV-19/USA/NY-NYCPHL-000108/2020|EPI_ISL_631600|2 .................... .................... ..................... .................... .................... ....................
Q 2135 hCoV-19/USA/NY-NYCPHL-000109/2020|EPI_ISL_631601|2 .................... .................... ..................... .................... .................... ....................
Q 2136 hCoV-19/USA/NY-NYCPHL-000110/2020|EPI_ISL_631602|2 .................... .................... ..................... .................... .................... ....................
Q 2137 hCoV-19/USA/NY-NYCPHL-000111/2020|EPI_ISL_631603|2 .................... .................... ..................... .................... .................... ....................
Q 2138 hCoV-19/USA/NY-NYCPHL-000112/2020|EPI_ISL_631604|2 .................... .................... ..................... .................... .................... ....................
Q 2139 hCoV-19/USA/NY-NYCPHL-000113/2020|EPI_ISL_631605|2 .................... .................... ..................... .................... .................... ....................
Q 2140 hCoV-19/USA/NY-NYCPHL-000114/2020|EPI_ISL_631606|2 .................... ..............C..... ..................... .................... .................... ....................
Q 2141 hCoV-19/USA/NY-NYCPHL-000115/2020|EPI_ISL_631607|2 .................... .................... ..................... .................... .................... ....................
Q 2142 hCoV-19/USA/NY-NYCPHL-000116/2020|EPI_ISL_631608|2 .................... .................... ..................... .................... .................... ....................
Q 2143 hCoV-19/USA/NY-NYCPHL-000117/2020|EPI_ISL_631609|2 .................... .................... ..................... .................... .................... ....................
Q 2144 hCoV-19/USA/NY-NYCPHL-000119/2020|EPI_ISL_631610|2 .................... .................... ..................... .................... .................... ....................
Q 2145 hCoV-19/USA/NY-NYCPHL-000121/2020|EPI_ISL_631612|2 .................... .................... ..................... .................... .................... ....................
Q 2146 hCoV-19/USA/NY-NYCPHL-000122/2020|EPI_ISL_631613|2 .................... .................... ..................... .................... .................... ....................
Q 2147 hCoV-19/USA/NY-NYCPHL-000124/2020|EPI_ISL_631615|2 .................... .................... ..................... .................... .................... ....................
Q 2148 hCoV-19/USA/NY-NYCPHL-000125/2020|EPI_ISL_631616|2 .................... .................... ..................... .................... .................... ....................
Q 2149 hCoV-19/USA/NY-NYCPHL-000126/2020|EPI_ISL_631617|2 .................... .................... ..................... .................... .................... ....................
Q 2150 hCoV-19/USA/NY-NYCPHL-000127/2020|EPI_ISL_631618|2 .................... .................... ..................... .................... .................... ....................
Q 2151 hCoV-19/USA/NY-NYCPHL-000128/2020|EPI_ISL_631619|2 .................... .................... ..................... .................... .................... ....................
Q 2152 hCoV-19/USA/NY-NYCPHL-000129/2020|EPI_ISL_631620|2 .................... .................... ..................... .................... .................... ....................
Q 2153 hCoV-19/USA/NY-NYCPHL-000130/2020|EPI_ISL_631621|2 .................... .................... ..................... .................... .................... ....................
Q 2154 hCoV-19/USA/NY-NYCPHL-000132/2020|EPI_ISL_631623|2 .................... .................... ..................... .................... .................... ....................
Q 2155 hCoV-19/USA/NY-NYCPHL-000133/2020|EPI_ISL_631624|2 .................... .................... ..................... .................... .................... ....................
Q 2156 hCoV-19/USA/NY-NYCPHL-000134/2020|EPI_ISL_631625|2 .................... .................... ..................... .................... .................... ....................
Q 2157 hCoV-19/USA/NY-NYCPHL-000135/2020|EPI_ISL_631626|2 .................... .................... ..................... .................... .................... ....................
Q 2158 hCoV-19/USA/NY-NYCPHL-000136/2020|EPI_ISL_631627|2 .................... .................... ..................... .................... .................... ....................
Q 2159 hCoV-19/USA/NY-NYCPHL-000137/2020|EPI_ISL_631628|2 .................... .................... ..................... .................... .................... ....................
Q 2160 hCoV-19/USA/NY-NYCPHL-000138/2020|EPI_ISL_631629|2 .................... .................... ..................... .................... .................... ....................
Q 2161 hCoV-19/USA/NY-NYCPHL-000139/2020|EPI_ISL_631630|2 .................... .................... ..................... .................... .................... ....................

Q 2162 hCoV-19/USA/NY-NYCPHL-000140/2020|EPI_ISL_631631|2 ···················· ···················· ···················· ···················· ···················· ····················
Q 2163 hCoV-19/USA/NY-NYCPHL-000141/2020|EPI_ISL_631632|2 ···················· ···················· ···················· ···················· ···················· ····················
Q 2164 hCoV-19/USA/NY-NYCPHL-000142/2020|EPI_ISL_631633|2 ···················· ···················· ···················· ···················· ···················· ····················
Q 2165 hCoV-19/USA/NY-NYCPHL-000143/2020|EPI_ISL_631634|2 ···················· ···················· ···················· ···················· ···················· ····················
Q 2166 hCoV-19/USA/NY-NYCPHL-000145/2020|EPI_ISL_631636|2 ···················· ···················· ···················· ···················· ···················· ····················
Q 2167 hCoV-19/USA/NY-NYCPHL-000146/2020|EPI_ISL_631637|2 ···················· ···················· ···················· ···················· ···················· ····················
Q 2168 hCoV-19/USA/WI-UW-1885/2020|EPI_ISL_632898|2020-09 ···················· ···················· ···················· ···················· ···················· ····················
Q 2169 hCoV-19/Bahrain/920457301_S8/2020|EPI_ISL_632899|2 ···················· ···················· ···················· ···················· ···················· ····················
Q 2170 hCoV-19/Bahrain/920457371_S3/2020|EPI_ISL_632900|2 ···················· ···················· ···················· ···················· ···················· ····················
Q 2171 hCoV-19/Bahrain/920457374_S4/2020|EPI_ISL_632901|2 ···················· ···················· ···················· ···················· ···················· ····················
Q 2172 hCoV-19/Bahrain/920458273_S24/2020|EPI_ISL_632902| ···················· ···················· ···················· ···················· ···················· ····················
Q 2173 hCoV-19/Bahrain/340862225_S35/2020|EPI_ISL_632903| ···················· ···················· ···················· ···················· ···················· ····················
Q 2174 hCoV-19/Bahrain/340798279/2020|EPI_ISL_632904|2020 ···················· ···················· ···················· ···················· ···················· ····················
Q 2175 hCoV-19/Bahrain/340861785/2020|EPI_ISL_632905|2020 ···················· ···················· ···················· ···················· ···················· ····················
Q 2176 hCoV-19/Bahrain/530223991_S30_L001/2020|EPI_ISL_63 ···················· ···················· ···················· ···················· ···················· ····················
Q 2177 hCoV-19/Bahrain/340862192_S34_L001/2020|EPI_ISL_63 ···················· ···················· ···················· ···················· ···················· ····················
Q 2178 hCoV-19/Pakistan/KP-RMI-01/2020|EPI_ISL_632908|202 ···················· ···················· ···················· ···················· ···················· ····················
Q 2179 hCoV-19/Canada/MB-NML-577/2020|EPI_ISL_632909|2020 ···················· ···················· ···················· ···················· ···················· ····················
Q 2180 hCoV-19/Canada/MB-NML-601/2020|EPI_ISL_632910|2020 ···················· ···················· ···················· ···················· ···················· ····················
Q 2181 hCoV-19/Canada/MB-NML-602/2020|EPI_ISL_632911|2020 ···················· ···················· ···················· ···················· ···················· ····················
Q 2182 hCoV-19/Canada/MB-NML-606/2020|EPI_ISL_632912|2020 ···················· ···················· ···················· ···················· ···················· ····················
Q 2183 hCoV-19/Canada/MB-NML-608/2020|EPI_ISL_632913|2020 ···················· ···················· ···················· ···················· ···················· ····················
Q 2184 hCoV-19/Canada/MB-NML-610/2020|EPI_ISL_632914|2020 ···················· ···················· ···················· ···················· ···················· ····················
Q 2185 hCoV-19/Canada/MB-NML-613/2020|EPI_ISL_632915|2020 ···················· ···················· ···················· ···················· ···················· ····················
Q 2186 hCoV-19/Canada/MB-NML-614/2020|EPI_ISL_632916|2020 ···················· ···················· ···················· ···················· ···················· ····················
Q 2187 hCoV-19/Canada/MB-NML-620/2020|EPI_ISL_632917|2020 ···················· ···················· ···················· ···················· ···················· ····················
Q 2188 hCoV-19/Canada/MB-NML-633/2020|EPI_ISL_632918|2020 ···················· ···················· ···················· ···················· ···················· ····················
Q 2189 hCoV-19/Canada/MB-NML-634/2020|EPI_ISL_632919|2020 ···················· ···················· ···················· ···················· ···················· ····················
Q 2190 hCoV-19/Canada/MB-NML-638/2020|EPI_ISL_632920|2020 ···················· ···················· ···················· ···················· ···················· ····················
Q 2191 hCoV-19/Canada/MB-NML-639/2020|EPI_ISL_632921|2020 ···················· ···················· ···················· ···················· ···················· ····················
Q 2192 hCoV-19/Canada/MB-NML-643/2020|EPI_ISL_632922|2020 ···················· ···················· ···················· ···················· ···················· ····················
Q 2193 hCoV-19/Canada/MB-NML-644/2020|EPI_ISL_632923|2020 ···················· ···················· ···················· ···················· ···················· ····················
Q 2194 hCoV-19/Canada/MB-NML-645/2020|EPI_ISL_632924|2020 ···················· ···················· ···················· ···················· ···················· ····················
Q 2195 hCoV-19/Canada/MB-NML-646/2020|EPI_ISL_632925|2020 ···················· ···················· ···················· ···················· ···················· ····················
Q 2196 hCoV-19/Canada/MB-NML-647/2020|EPI_ISL_632926|2020 ···················· ···················· ···················· ···················· ···················· ····················
Q 2197 hCoV-19/Canada/MB-NML-648/2020|EPI_ISL_632927|2020 ···················· ···················· ···················· ···················· ···················· ····················
Q 2198 hCoV-19/Canada/MB-NML-649/2020|EPI_ISL_632928|2020 ···················· ···················· ···················· ···················· ···················· ····················
Q 2199 hCoV-19/Canada/MB-NML-650/2020|EPI_ISL_632929|2020 ···················· ···················· ···················· ···················· ···················· ····················
Q 2200 hCoV-19/Canada/MB-NML-651/2020|EPI_ISL_632930|2020 ···················· ···················· ···················· ···················· ···················· ····················
Q 2201 hCoV-19/Canada/MB-NML-652/2020|EPI_ISL_632931|2020 ···················· ···················· ···················· ···················· ···················· ····················
Q 2202 hCoV-19/Canada/MB-NML-653/2020|EPI_ISL_632932|2020 ···················· ···················· ···················· ···················· ···················· ····················
Q 2203 hCoV-19/Canada/MB-NML-654/2020|EPI_ISL_632933|2020 ···················· ···················· ···················· ···················· ···················· ····················
Q 2204 hCoV-19/Yunnan/02/2020|EPI_ISL_632934|2020-09-13 ···················· ·A·················· ···················· ···················· ···················· ··A·················
Q 2205 hCoV-19/Indonesia/YO-UGM-10007/2020|EPI_ISL_632936 ·······T············ ···················· ···················· ···················· ···················· ····················
Q 2206 hCoV-19/Indonesia/YO-UGM-48651/2020|EPI_ISL_632937 ···················· ···················· ···················· ···················· ···················· ····················
Q 2207 hCoV-19/USA/NY-NYCPHL-000866/2020|EPI_ISL_632939|2 ···················· ···················· ···················· ···················· ···················· ····················
Q 2208 hCoV-19/USA/NY-NYCPHL-000867/2020|EPI_ISL_632940|2 ···················· ···················· ···················· ···················· ···················· ····················
Q 2209 hCoV-19/USA/NY-NYCPHL-000868/2020|EPI_ISL_632941|2 ···················· ···················· ···················· ···················· ···················· ····················
Q 2210 hCoV-19/USA/NY-NYCPHL-000869/2020|EPI_ISL_632942|2 ···················· ···················· ···················· ···················· ···················· ····················
Q 2211 hCoV-19/USA/NY-NYCPHL-000872/2020|EPI_ISL_632944|2 ···················· ···················· ···················· ···················· ···················· ····················
Q 2212 hCoV-19/USA/NY-NYCPHL-000873/2020|EPI_ISL_632945|2 ···················· ···················· ···················· ···················· ···················· ····················
Q 2213 hCoV-19/USA/NY-NYCPHL-000874/2020|EPI_ISL_632946|2 ···················· ···················· ···················· ···················· ···················· ····················
Q 2214 hCoV-19/USA/NY-NYCPHL-000875/2020|EPI_ISL_632947|2 ···················· ···················· ···················· ···················· ···················· ····················
Q 2215 hCoV-19/USA/NY-NYCPHL-000876/2020|EPI_ISL_632948|2 ···················· ···················· ···················· ···················· ···················· ····················
Q 2216 hCoV-19/USA/NY-NYCPHL-000877/2020|EPI_ISL_632949|2 ···················· ···················· ···················· ···················· ···················· ····················
Q 2217 hCoV-19/USA/NY-NYCPHL-000878/2020|EPI_ISL_632950|2 ···················· ···················· ···················· ···················· ···················· ····················
Q 2218 hCoV-19/USA/NY-NYCPHL-000879/2020|EPI_ISL_632951|2 ···················· ···················· ···················· ···················· ···················· ····················
Q 2219 hCoV-19/USA/NY-NYCPHL-000880/2020|EPI_ISL_632952|2 ···················· ···················· ···················· ···················· ···················· ····················
Q 2220 hCoV-19/USA/NY-NYCPHL-000881/2020|EPI_ISL_632953|2 ···················· ···················· ···················· ···················· ···················· ····················
Q 2221 hCoV-19/USA/NY-NYCPHL-000882/2020|EPI_ISL_632954|2 ···················· ···················· ···················· ···················· ···················· ····················
Q 2222 hCoV-19/USA/NY-NYCPHL-000883/2020|EPI_ISL_632955|2 ···················· ···················· ···················· ···················· ···················· ····················
Q 2223 hCoV-19/USA/NY-NYCPHL-000884/2020|EPI_ISL_632956|2 ···················· ···················· ···················· ···················· ···················· ····················
Q 2224 hCoV-19/USA/NY-NYCPHL-000885/2020|EPI_ISL_632957|2 ···················· ···················· ···················· ···················· ···················· ····················
Q 2225 hCoV-19/USA/NY-NYCPHL-000886/2020|EPI_ISL_632958|2 ···················· ···················· ···················· ···················· ···················· ····················
Q 2226 hCoV-19/USA/NY-NYCPHL-000887/2020|EPI_ISL_632959|2 ···················· ···················· ···················· ···················· ···················· ····················
Q 2227 hCoV-19/USA/NY-NYCPHL-000889/2020|EPI_ISL_632960|2 ···················· ···················· ···················· ···················· ···················· ····················
Q 2228 hCoV-19/USA/NY-NYCPHL-000891/2020|EPI_ISL_632961|2 ···················· ···················· ···················· ···················· ···················· ····················
Q 2229 hCoV-19/USA/NY-NYCPHL-000892/2020|EPI_ISL_632962|2 ···················· ···················· ···················· ···················· ···················· ····················
Q 2230 hCoV-19/USA/NY-NYCPHL-000894/2020|EPI_ISL_632963|2 ···················· ···················· ···················· ···················· ···················· ····················
Q 2231 hCoV-19/USA/NY-NYCPHL-000898/2020|EPI_ISL_632964|2 ···················· ···················· ···················· ···················· ···················· ····················
Q 2232 hCoV-19/USA/NY-NYCPHL-000899/2020|EPI_ISL_632965|2 ···················· ···················· ···················· ···················· ···················· ····················
Q 2233 hCoV-19/USA/NY-NYCPHL-000905/2020|EPI_ISL_632966|2 ···················· ···················· ···················· ···················· ···················· ····················
Q 2234 hCoV-19/USA/NY-NYCPHL-000906/2020|EPI_ISL_632967|2 ···················· ···················· ···················· ···················· ···················· ····················
Q 2235 hCoV-19/USA/NY-NYCPHL-000907/2020|EPI_ISL_632968|2 ···················· ···················· ···················· ···················· ···················· ····················
Q 2236 hCoV-19/USA/NY-NYCPHL-000908/2020|EPI_ISL_632969|2 ···················· ···················· ···················· ···················· ···················· ····················
Q 2237 hCoV-19/USA/NY-NYCPHL-000909/2020|EPI_ISL_632970|2 ···················· ···················· ···················· ···················· ···················· ····················
Q 2238 hCoV-19/USA/NY-NYCPHL-000910/2020|EPI_ISL_632971|2 ···················· ···················· ···················· ···················· ···················· ····················
Q 2239 hCoV-19/USA/NY-NYCPHL-000911/2020|EPI_ISL_632972|2 ···················· ···················· ···················· ···················· ···················· ····················
Q 2240 hCoV-19/USA/NY-NYCPHL-000913/2020|EPI_ISL_632974|2 ···················· ···················· ···················· ···················· ···················· ····················
Q 2241 hCoV-19/Norway/3735/2020|EPI_ISL_635109|2020-09-09 ···················· ···················· ···················· ···················· ···················· ····················

| | | | | | | | | |
|---|---|---|---|---|---|---|---|---|
| Q | 2242 hCoV-19/Norway/3627/2020\|EPI_ISL_635110\|2020-09-25 | ···················· | ·A·················· | ···················· | ···················· | ···················· | ··A················· | |
| Q | 2243 hCoV-19/Norway/3628/2020\|EPI_ISL_635111\|2020-09-25 | ···················· | ·A·················· | ···················· | ···················· | ···················· | ··A················· | |
| Q | 2244 hCoV-19/Norway/3908/2020\|EPI_ISL_635112\|2020-09-21 | ···················· | ···················· | ···················· | ···················· | ···················· | ···················· | |
| Q | 2245 hCoV-19/Norway/3550/2020\|EPI_ISL_635113\|2020-08-06 | ···················· | ·A·················· | ···················· | ···················· | ···················· | ··A················· | |
| Q | 2246 hCoV-19/Norway/3563/2020\|EPI_ISL_635114\|2020-08-06 | ···················· | ·A·················· | ···················· | ···················· | ···················· | ··A················· | |
| Q | 2247 hCoV-19/Norway/3570/2020\|EPI_ISL_635115\|2020-07-31 | ···················· | ·A·················· | ···················· | ···················· | ···················· | ··A················· | |
| Q | 2248 hCoV-19/Norway/3620/2020\|EPI_ISL_635116\|2020-05-22 | ···················· | ···················· | ···················· | ···················· | ···················· | ···················· | |
| Q | 2249 hCoV-19/Norway/3573/2020\|EPI_ISL_635117\|2020-07-31 | ···················· | ·A·················· | ···················· | ···················· | ···················· | ··A················· | |
| Q | 2250 hCoV-19/Norway/3575/2020\|EPI_ISL_635118\|2020-07-31 | ···················· | ·A·················· | ···················· | ···················· | ···················· | ··A················· | |
| Q | 2251 hCoV-19/Norway/3713/2020\|EPI_ISL_635119\|2020-09-18 | ···················· | ···················· | ···················· | ···················· | ···················· | ···················· | |
| Q | 2252 hCoV-19/Norway/3920/2020\|EPI_ISL_635123\|2020-09-28 | ···················· | ···················· | ···················· | ···················· | ···················· | ···················· | |
| Q | 2253 hCoV-19/Norway/4040/2020\|EPI_ISL_635124\|2020-10-02 | ···················· | ···················· | ···················· | ···················· | ···················· | ···················· | |
| Q | 2254 hCoV-19/Norway/4047/2020\|EPI_ISL_635125\|2020-10-02 | ···················· | ···················· | ···················· | ···················· | ···················· | ···················· | |
| Q | 2255 hCoV-19/Norway/4055/2020\|EPI_ISL_635126\|2020-09-30 | ···················· | ···················· | ···················· | ···················· | ···················· | ···················· | |
| Q | 2256 hCoV-19/Norway/4061/2020\|EPI_ISL_635127\|2020-09-29 | ···················· | ···················· | ···················· | ···················· | ···················· | ···················· | |
| Q | 2257 hCoV-19/Norway/4100/2020\|EPI_ISL_635128\|2020-10-05 | ···················· | ···················· | ···················· | ···················· | ···················· | ···················· | |
| Q | 2258 hCoV-19/Norway/4123/2020\|EPI_ISL_635129\|2020-10-02 | ···················· | ···················· | ···················· | ···················· | ···················· | ···················· | |
| Q | 2259 hCoV-19/Norway/4612/2020\|EPI_ISL_635170\|2020-10-07 | ···················· | ···················· | ···················· | ···················· | ···················· | ···················· | |
| Q | 2260 hCoV-19/Norway/4545/2020\|EPI_ISL_635171\|2020-10-07 | ···················· | ···················· | ···················· | ···················· | ···················· | ···················· | |
| Q | 2261 hCoV-19/Norway/4125/2020\|EPI_ISL_635172\|2020-10-04 | ···················· | ···················· | ···················· | ···················· | ···················· | ···················· | |
| Q | 2262 hCoV-19/Norway/4620/2020\|EPI_ISL_635173\|2020-10-06 | ···················· | ···················· | ···················· | ···················· | ···················· | ···················· | |
| Q | 2263 hCoV-19/Norway/4630/2020\|EPI_ISL_635174\|2020-10-18 | ···················· | ···················· | ···················· | ···················· | ···················· | ···················· | |
| Q | 2264 hCoV-19/Norway/4633/2020\|EPI_ISL_635175\|2020-10-18 | ···················· | ···················· | ···················· | ···················· | ···················· | ···················· | |
| Q | 2265 hCoV-19/Norway/4621/2020\|EPI_ISL_635176\|2020-10-06 | ···················· | ···················· | ···················· | ···················· | ···················· | ···················· | |
| Q | 2266 hCoV-19/Norway/4614/2020\|EPI_ISL_635177\|2020-10-02 | ···················· | ···················· | ···················· | ···················· | ···················· | ···················· | |
| Q | 2267 hCoV-19/Norway/4688/2020\|EPI_ISL_635178\|2020-10-22 | ···················· | ···················· | ···················· | ···················· | ···T················ | ···················· | |
| Q | 2268 hCoV-19/Norway/4696/2020\|EPI_ISL_635179\|2020-10-14 | ···················· | ···················· | ···················· | ···················· | ···················· | ···················· | |
| Q | 2269 hCoV-19/Norway/4699/2020\|EPI_ISL_635180\|2020-10-16 | ···················· | ···················· | ···················· | ···················· | ···················· | ···················· | |
| Q | 2270 hCoV-19/Norway/4700/2020\|EPI_ISL_635181\|2020-10-19 | ···················· | ···················· | ···················· | ···················· | ···················· | ···················· | |
| Q | 2271 hCoV-19/Norway/4336/2020\|EPI_ISL_635182\|2020-10-12 | ···················· | ·A·················· | ···················· | ···················· | ···················· | ···················· | |
| Q | 2272 hCoV-19/Norway/4314/2020\|EPI_ISL_635183\|2020-10-06 | ···················· | ···················· | ···················· | ···················· | ···················· | ···················· | |
| Q | 2273 hCoV-19/Norway/4108/2020\|EPI_ISL_635184\|2020-09-23 | ···················· | ···················· | ···················· | ···················· | ···················· | ···················· | |
| Q | 2274 hCoV-19/Norway/3621/2020\|EPI_ISL_635185\|2020-09-20 | ···················· | ···················· | ···················· | ···················· | ···················· | ···················· | |
| Q | 2275 hCoV-19/Norway/3584/2020\|EPI_ISL_635186\|2020-07-31 | ···················· | ·A·················· | ···················· | ···················· | ···················· | ··A················· | |
| Q | 2276 hCoV-19/Norway/3529/2020\|EPI_ISL_635187\|2020-09-22 | ···················· | ···················· | ···················· | ···················· | ···················· | ···················· | |
| Q | 2277 hCoV-19/Norway/3646/2020\|EPI_ISL_635188\|2020-09-25 | ···················· | ···················· | ···················· | ···················· | ···················· | ···················· | |
| Q | 2278 hCoV-19/Norway/4282/2020\|EPI_ISL_635189\|2020-10-11 | ···················· | ···················· | ···················· | ···················· | ···················· | ···················· | |
| Q | 2279 hCoV-19/Norway/4308/2020\|EPI_ISL_635190\|2020-10-06 | ···················· | ···················· | ···················· | ···················· | ···················· | ···················· | |
| Q | 2280 hCoV-19/Norway/4594/2020\|EPI_ISL_635191\|2020-10-12 | ···················· | ···················· | ···················· | ···················· | ···················· | ···················· | |
| Q | 2281 hCoV-19/Norway/4616/2020\|EPI_ISL_635192\|2020-10-17 | ···················· | ···················· | ···················· | ···················· | ···················· | ···················· | |
| Q | 2282 hCoV-19/Norway/4628/2020\|EPI_ISL_635193\|2020-10-17 | ···················· | ···················· | ···················· | ···················· | ···················· | ···················· | |
| Q | 2283 hCoV-19/Norway/4637/2020\|EPI_ISL_635194\|2020-10-19 | ···················· | ···················· | ···················· | ···················· | ···················· | ···················· | |
| Q | 2284 hCoV-19/Norway/4547/2020\|EPI_ISL_635195\|2020-10-15 | ···················· | ···················· | ···················· | ···················· | ···················· | ···················· | |
| Q | 2285 hCoV-19/Norway/4555/2020\|EPI_ISL_635196\|2020-10-12 | ···················· | ···················· | ···················· | ···················· | ···················· | ···················· | |
| Q | 2286 hCoV-19/Norway/4693/2020\|EPI_ISL_635197\|2020-10-06 | ···················· | ·A·················· | ···················· | ···················· | ···················· | ··A················· | |
| Q | 2287 hCoV-19/Norway/4747/2020\|EPI_ISL_635198\|2020-10-19 | ···················· | ···················· | ···················· | ···················· | ···················· | ···················· | |
| Q | 2288 hCoV-19/Jordan/AM-ALSR-4080/2020\|EPI_ISL_636440\|20 | ···················· | ···················· | ···················· | ···················· | ···················· | ···················· | |
| Q | 2289 hCoV-19/Jordan/AM-ALSR-4081/2020\|EPI_ISL_636441\|20 | ···················· | ···················· | ···················· | ···················· | ···················· | ···················· | |
| Q | 2290 hCoV-19/Jordan/AM-ALSR-4082/2020\|EPI_ISL_636442\|20 | ···················· | ···················· | ···················· | ···················· | ···················· | ···················· | |
| Q | 2291 hCoV-19/Jordan/AM-ALSR-4083/2020\|EPI_ISL_636443\|20 | ···················· | ···················· | ···················· | ···················· | ···················· | ···················· | |
| Q | 2292 hCoV-19/Jordan/AM-ALSR-4084/2020\|EPI_ISL_636444\|20 | ···················· | ···················· | ···················· | ···················· | ···················· | ···················· | |
| Q | 2293 hCoV-19/Jordan/AM-ALSR-4085/2020\|EPI_ISL_636445\|20 | ···················· | ···················· | ···················· | ···················· | ···················· | ···················· | |
| Q | 2294 hCoV-19/Jordan/AM-ALSR-4086/2020\|EPI_ISL_636446\|20 | ···················· | ···················· | ···················· | ···················· | ···················· | ···················· | |
| Q | 2295 hCoV-19/Jordan/AM-ALSR-4087/2020\|EPI_ISL_636447\|20 | ···················· | ···················· | ···················· | ···················· | ···················· | ···················· | |
| Q | 2296 hCoV-19/Jordan/AM-ALSR-4088/2020\|EPI_ISL_636448\|20 | ···················· | ···················· | ···················· | ···················· | ···················· | ···················· | |
| Q | 2297 hCoV-19/Jordan/AM-ALSR-4089/2020\|EPI_ISL_636449\|20 | ···················· | ···················· | ···················· | ···················· | ···················· | ···················· | |
| Q | 2298 hCoV-19/Jordan/AM-ALSR-4090/2020\|EPI_ISL_636450\|20 | ···················· | ···················· | ···················· | ···················· | ···················· | ···················· | |
| Q | 2299 hCoV-19/Jordan/AM-ALSR-4091/2020\|EPI_ISL_636451\|20 | ···················· | ···················· | ···················· | ···················· | ···················· | ···················· | |
| Q | 2300 hCoV-19/Jordan/AM-ALSR-4092/2020\|EPI_ISL_636452\|20 | ···················· | ···················· | ···················· | ···················· | ···················· | ···················· | |
| Q | 2301 hCoV-19/Jordan/AM-ALSR-4093/2020\|EPI_ISL_636453\|20 | ···················· | ···················· | ···················· | ···················· | ···················· | ···················· | |
| Q | 2302 hCoV-19/Jordan/AM-ALSR-4096/2020\|EPI_ISL_636456\|20 | ···················· | ···················· | ···················· | ···················· | ···················· | ···················· | |
| Q | 2303 hCoV-19/Jordan/AM-ALSR-4097/2020\|EPI_ISL_636457\|20 | ···················· | ···················· | ···················· | ···················· | ···················· | ···················· | |
| Q | 2304 hCoV-19/Jordan/AM-ALSR-4098/2020\|EPI_ISL_636458\|20 | ···················· | ···················· | ···················· | ···················· | ···················· | ···················· | |
| Q | 2305 hCoV-19/Scotland/CVR5229/2020\|EPI_ISL_627400\|2020- | ···················· | ···················· | ···················· | ···················· | ···················· | ···················· | |
| Q | 2306 hCoV-19/Scotland/CVR5231/2020\|EPI_ISL_627401\|2020- | ···················· | ···················· | ···················· | ···················· | ···················· | ···················· | |
| Q | 2307 hCoV-19/Scotland/CVR5233/2020\|EPI_ISL_627402\|2020- | ···················· | ···················· | ···················· | ···················· | ···················· | ···················· | |
| Q | 2308 hCoV-19/Scotland/CVR5234/2020\|EPI_ISL_627403\|2020- | ···················· | ···················· | ···················· | ···················· | ···················· | NN·····N············ | 1 |
| Q | 2309 hCoV-19/Scotland/CVR5236/2020\|EPI_ISL_627404\|2020- | ···················· | ···················· | ···················· | ···················· | ···················· | ···················· | |
| Q | 2310 hCoV-19/Scotland/CVR5238/2020\|EPI_ISL_627405\|2020- | ···················· | ···················· | ···················· | ···················· | ···················· | ···················· | |
| Q | 2311 hCoV-19/Scotland/CVR5239/2020\|EPI_ISL_627406\|2020- | ···················· | ···················· | ···················· | ···················· | ···················· | ···················· | |
| Q | 2312 hCoV-19/Scotland/CVR5241/2020\|EPI_ISL_627407\|2020- | ···················· | ···················· | ···················· | ···················· | ···················· | ···················· | |
| Q | 2313 hCoV-19/Scotland/CVR5242/2020\|EPI_ISL_627408\|2020- | ···················· | ···················· | ···················· | ···················· | ···················· | ···················· | |
| Q | 2314 hCoV-19/Scotland/CVR5245/2020\|EPI_ISL_627409\|2020- | ···················· | ···················· | ···················· | ···················· | ···················· | ···················· | |
| Q | 2315 hCoV-19/Scotland/CVR5250/2020\|EPI_ISL_627412\|2020- | ···················· | ···················· | ···················· | ···················· | ···················· | ···················· | |
| Q | 2316 hCoV-19/Scotland/CVR5251/2020\|EPI_ISL_627413\|2020- | ···················· | ···················· | ···················· | ···················· | ···················· | ···················· | |
| Q | 2317 hCoV-19/Scotland/CVR5252/2020\|EPI_ISL_627414\|2020- | ···················· | ···················· | ···················· | ···················· | ···················· | ···················· | |
| Q | 2318 hCoV-19/Scotland/CVR5253/2020\|EPI_ISL_627415\|2020- | ···················· | ···················· | ···················· | ···················· | ···················· | ···················· | |
| Q | 2319 hCoV-19/Scotland/CVR5256/2020\|EPI_ISL_627418\|2020- | ···················· | ···················· | ···················· | ···················· | ···················· | ···················· | |
| Q | 2320 hCoV-19/Scotland/CVR5260/2020\|EPI_ISL_627420\|2020- | ···················· | ···················· | ···················· | ···················· | ···················· | ···················· | |
| Q | 2321 hCoV-19/Scotland/CVR5262/2020\|EPI_ISL_627421\|2020- | ···················· | ···················· | ···················· | ···················· | ···················· | ···················· | |

Q 2322 hCoV-19/Scotland/CVR5263/2020|EPI_ISL_627422|2020- ···················· ···················· ···················· ···················· ···················· ····················
Q 2323 hCoV-19/Scotland/CVR5266/2020|EPI_ISL_627423|2020- ···················· ···················· ···················· ···················· ···················· ····················
Q 2324 hCoV-19/England/EXET-13860C/2020|EPI_ISL_627424|20 ···················· ···················· ···················· ···················· ···················· ····················
Q 2325 hCoV-19/England/EXET-1386EE/2020|EPI_ISL_627425|20 ···················· ···················· ···················· ···················· ···················· ····················
Q 2326 hCoV-19/England/EXET-138736/2020|EPI_ISL_627426|20 ···················· ···················· ···················· ···················· ···················· ····················
Q 2327 hCoV-19/England/EXET-138790/2020|EPI_ISL_627427|20 ···················· ···················· ···················· ···················· ···················· ····················
Q 2328 hCoV-19/England/EXET-1387AF/2020|EPI_ISL_627428|20 ···················· ···················· ···················· ···················· ···················· ····················
Q 2329 hCoV-19/England/EXET-138851/2020|EPI_ISL_627429|20 ···················· ···················· ···················· ···················· ···················· ····················
Q 2330 hCoV-19/England/MILK-AC8653/2020|EPI_ISL_629210|20 ···················· ···················· ···················· ···················· ···················· ····················
Q 2331 hCoV-19/England/MILK-AC842C/2020|EPI_ISL_629211|20 ···················· ···················· ···················· ···················· ···················· ····················
Q 2332 hCoV-19/England/MILK-AC84A4/2020|EPI_ISL_629212|20 ···················· ···················· ···················· ···················· ···················· ····················
Q 2333 hCoV-19/England/MILK-ACED3A/2020|EPI_ISL_629213|20 ···················· ···················· ···················· ···················· ···················· ····················
Q 2334 hCoV-19/England/MILK-ACBDB5/2020|EPI_ISL_629214|20 ···················· ···················· ···················· ···················· ···················· ····················
Q 2335 hCoV-19/England/MILK-AC85CF/2020|EPI_ISL_629215|20 ···················· ···················· ···················· ···················· ·····T·············· ····················
Q 2336 hCoV-19/England/MILK-ACBF28/2020|EPI_ISL_629216|20 ···················· ···················· ···················· ···················· ···················· ····················
Q 2337 hCoV-19/England/MILK-ACBADC/2020|EPI_ISL_629218|20 ···················· ···················· ···················· ···················· ···················· ····················
Q 2338 hCoV-19/England/MILK-ACC200/2020|EPI_ISL_629220|20 ···················· ···················· ···················· ···················· ···················· ····················
Q 2339 hCoV-19/England/MILK-ACC21F/2020|EPI_ISL_629221|20 ···················· ···················· ···················· ···················· ···················· ····················
Q 2340 hCoV-19/England/MILK-ACBB9D/2020|EPI_ISL_629222|20 ···················· ···················· ···················· ···················· ···················· ····················
Q 2341 hCoV-19/England/MILK-ACC121/2020|EPI_ISL_629223|20 ···················· ···················· ···················· ···················· ···················· ····················
Q 2342 hCoV-19/England/MILK-ACC015/2020|EPI_ISL_629224|20 ···················· ···················· ···················· ···················· ···················· ····················
Q 2343 hCoV-19/England/MILK-ACBDA6/2020|EPI_ISL_629225|20 ······C············· ···················· ···················· ···················· ···················· ····················
Q 2344 hCoV-19/England/MILK-AC95EC/2020|EPI_ISL_629226|20 ···················· ·A·················· ···················· ···················· ···················· ··A·················
Q 2345 hCoV-19/England/MILK-AC7CF8/2020|EPI_ISL_629227|20 ··········T········· ···················· ···················· ···················· ···················· ····················
Q 2346 hCoV-19/England/MILK-ACC130/2020|EPI_ISL_629228|20 ···················· ···················· ···················· ···················· ···················· ····················
Q 2347 hCoV-19/England/MILK-ACBFFB/2020|EPI_ISL_629230|20 ···················· ···················· ···················· ···················· ···················· ····················
Q 2348 hCoV-19/England/MILK-ACC24C/2020|EPI_ISL_629231|20 ···················· ···················· ···················· ···················· ···················· ····················
Q 2349 hCoV-19/England/MILK-ACE4DB/2020|EPI_ISL_629232|20 ···················· ···················· ···················· ···················· ···················· ····················
Q 2350 hCoV-19/England/MILK-ACECD3/2020|EPI_ISL_629233|20 ···················· ···················· ···················· ···················· ···················· ····················
Q 2351 hCoV-19/England/MILK-AC85B0/2020|EPI_ISL_629234|20 ···················· ···················· ···················· ···················· ···················· ····················
Q 2352 hCoV-19/England/MILK-ACBD5B/2020|EPI_ISL_629235|20 ···················· ···················· ···················· ···················· ···················· ····················
Q 2353 hCoV-19/England/MILK-AC8459/2020|EPI_ISL_629236|20 ···················· ···················· ···················· ···················· ···················· ····················
Q 2354 hCoV-19/England/MILK-AC7CCB/2020|EPI_ISL_629237|20 ···················· ···················· ···················· ···················· ···················· ····················
Q 2355 hCoV-19/England/MILK-ACBED0/2020|EPI_ISL_629238|20 ···················· ···················· ···················· ···················· ···················· ····················
Q 2356 hCoV-19/England/MILK-AC83A7/2020|EPI_ISL_629239|20 ···················· ···················· ···················· ···················· ···················· ····················
Q 2357 hCoV-19/England/MILK-ACBD2E/2020|EPI_ISL_629240|20 ···················· ···················· ···················· ···················· ···················· ····················
Q 2358 hCoV-19/England/MILK-AC86AE/2020|EPI_ISL_629242|20 ···················· ·A·················· ···················· ···················· ···················· ··A·················
Q 2359 hCoV-19/England/MILK-ACEBE5/2020|EPI_ISL_629243|20 ···················· ···················· ···················· ···················· ···················· ····················
Q 2360 hCoV-19/England/MILK-ACC0E8/2020|EPI_ISL_629244|20 ···················· ·A·················· ···················· ···················· ···················· ··A·················
Q 2361 hCoV-19/England/MILK-ACE8FD/2020|EPI_ISL_629246|20 ···················· ···················· ···················· ···················· ···················· ····················
Q 2362 hCoV-19/England/MILK-ACC18B/2020|EPI_ISL_629247|20 ···················· ···················· ···················· ···················· ···················· ····················
Q 2363 hCoV-19/England/MILK-ACC886/2020|EPI_ISL_629248|20 ···················· ···················· ···················· ···················· ·····T·············· ····················
Q 2364 hCoV-19/England/MILK-ACC30D/2020|EPI_ISL_629249|20 ···················· ···················· ···················· ···················· ·····T·············· ····················
Q 2365 hCoV-19/England/MILK-AC8468/2020|EPI_ISL_629253|20 ···················· ···················· ···················· ···················· ···················· ····················
Q 2366 hCoV-19/England/MILK-AC92C7/2020|EPI_ISL_629256|20 ···················· ···················· ···················· ···················· ···················· ····················
Q 2367 hCoV-19/England/MILK-ACBF46/2020|EPI_ISL_629257|20 ···················· ·A·················· ···················· ···················· ···················· ··A·················
Q 2368 hCoV-19/England/MILK-AC833E/2020|EPI_ISL_629258|20 ···················· ···················· ···················· ···················· ···················· ····················
Q 2369 hCoV-19/England/MILK-ACC3A3/2020|EPI_ISL_629259|20 ···················· ···················· ···················· ···················· ·····T·············· ····················
Q 2370 hCoV-19/England/MILK-ACE3CF/2020|EPI_ISL_629270|20 ···················· ···················· ···················· ···················· ·····T·············· ····················
Q 2371 hCoV-19/England/MILK-ACE471/2020|EPI_ISL_629271|20 ···················· ···················· ···················· ···················· ···················· ····················
Q 2372 hCoV-19/England/MILK-AC82C8/2020|EPI_ISL_629273|20 ···················· ···················· ···················· ···················· ···················· ····················
Q 2373 hCoV-19/England/MILK-ACE4EA/2020|EPI_ISL_629274|20 ···················· ···················· ···················· ···················· ···················· ····················
Q 2374 hCoV-19/England/MILK-AC927C/2020|EPI_ISL_629275|20 ···················· ·A·················· ···················· ···················· ···················· ··A·················
Q 2375 hCoV-19/England/MILK-ACBFEC/2020|EPI_ISL_629278|20 ···················· ···················· ···················· ···················· ·····T·············· ····················
Q 2376 hCoV-19/England/MILK-ACE954/2020|EPI_ISL_629279|20 ···················· ···················· ···················· ···················· ···················· ····················
Q 2377 hCoV-19/England/MILK-ACBCF4/2020|EPI_ISL_629280|20 ···················· ···················· ···················· ···················· ···················· ····················
Q 2378 hCoV-19/England/MILK-ACEC3D/2020|EPI_ISL_629281|20 ···················· ···················· ···················· ···················· ·····T·············· ····················
Q 2379 hCoV-19/England/MILK-ACC297/2020|EPI_ISL_629282|20 ···················· ···················· ···················· ···················· ···················· ····················
Q 2380 hCoV-19/England/MILK-ACBEEF/2020|EPI_ISL_629283|20 ···················· ···················· ···················· ···················· ···················· ····················
Q 2381 hCoV-19/England/MILK-AC7D5F/2020|EPI_ISL_629284|20 ···················· ···················· ···················· ···················· ···················· ····················
Q 2382 hCoV-19/England/MILK-ACC3B2/2020|EPI_ISL_629285|20 ···················· ···················· ···················· ···················· ···················· ····················
Q 2383 hCoV-19/England/MILK-ACBFCE/2020|EPI_ISL_629286|20 ···················· ···················· ···················· ···················· ···················· ····················
Q 2384 hCoV-19/England/MILK-ACE22C/2020|EPI_ISL_629287|20 ···················· ···················· ···················· ···················· ···················· ····················
Q 2385 hCoV-19/England/MILK-ACECF1/2020|EPI_ISL_629288|20 ···················· ···················· ···················· ···················· ···················· ····················
Q 2386 hCoV-19/England/MILK-ACE3B0/2020|EPI_ISL_629289|20 ···················· ·A·················· ···················· ···················· ···················· ··A·················
Q 2387 hCoV-19/England/MILK-AC83D4/2020|EPI_ISL_629290|20 ···················· ···················· ···················· ···················· ·····T·············· ····················
Q 2388 hCoV-19/England/MILK-ACE8EE/2020|EPI_ISL_629291|20 ···················· ···················· ···················· ···················· ···················· ····················
Q 2389 hCoV-19/England/MILK-ACE31A/2020|EPI_ISL_629292|20 ···················· ···················· ···················· ···················· ···················· ····················
Q 2390 hCoV-19/England/MILK-ACE8C0/2020|EPI_ISL_629293|20 ···················· ···················· ···················· ···················· ···················· ····················
Q 2391 hCoV-19/England/MILK-AC7D13/2020|EPI_ISL_629294|20 ···················· ···················· ···················· ···················· ···················· ····················
Q 2392 hCoV-19/England/MILK-ACE480/2020|EPI_ISL_629295|20 ···················· ···················· ···················· ···················· ···················· ····················
Q 2393 hCoV-19/England/MILK-ACBFBF/2020|EPI_ISL_629296|20 ···················· ···················· ···················· ···················· ·····T·············· ····················
Q 2394 hCoV-19/England/MILK-ACCA35/2020|EPI_ISL_629297|20 ···················· ···················· ···················· ···················· ·····T·············· ··············A·····
Q 2395 hCoV-19/England/MILK-AC9625/2020|EPI_ISL_629298|20 ···················· ···················· ···················· ···················· ···················· ····················
Q 2396 hCoV-19/England/MILK-AC844A/2020|EPI_ISL_629299|20 ···················· ···················· ···················· ···················· ···················· ····················
Q 2397 hCoV-19/England/MILK-ACDCC5/2020|EPI_ISL_630200|20 ···················· ···················· ···················· ···················· ···················· ····················
Q 2398 hCoV-19/England/MILK-ACDD2C/2020|EPI_ISL_630201|20 ···················· ···················· ···················· ···················· ···················· ····················
Q 2399 hCoV-19/England/MILK-ACCFCD/2020|EPI_ISL_630202|20 ···················· ···················· ···················· ···················· ···················· ····················
Q 2400 hCoV-19/England/MILK-ACE1F2/2020|EPI_ISL_630203|20 ···················· ···················· ···················· ···················· ·····T·············· ····················
Q 2401 hCoV-19/England/MILK-AC9A47/2020|EPI_ISL_630204|20 ···················· ···················· ···················· ···················· ···················· ····················

| | | | | | | | | |
|---|---|---|---|---|---|---|---|---|
| Q | 2402 | hCoV-19/England/MILK-AC7399/2020\|EPI_ISL_630206\|20 | ···················· | ·A·················· | ···················· | ···················· | ···················· | ···················· |
| Q | 2403 | hCoV-19/England/MILK-ACD8A3/2020\|EPI_ISL_630207\|20 | ···················· | ···················· | ···················· | ···················· | ···················· | ···················· |
| Q | 2404 | hCoV-19/England/MILK-AC740F/2020\|EPI_ISL_630208\|20 | ···················· | ···················· | ···················· | ···················· | ···················· | ···················· |
| Q | 2405 | hCoV-19/England/MILK-AC74E1/2020\|EPI_ISL_630209\|20 | ···················· | ···················· | ···················· | ···················· | ···················· | ···················· |
| Q | 2406 | hCoV-19/England/MILK-ACDCE3/2020\|EPI_ISL_630211\|20 | ···················· | ·A·················· | ···················· | ···················· | ···················· | ···A················ |
| Q | 2407 | hCoV-19/England/MILK-ACDBF5/2020\|EPI_ISL_630212\|20 | ···················· | ···················· | ···················· | ···················· | ···················· | ···················· |
| Q | 2408 | hCoV-19/England/MILK-AC933D/2020\|EPI_ISL_630213\|20 | ···················· | ···················· | ···················· | ···················· | ···················· | ···················· |
| Q | 2409 | hCoV-19/England/MILK-ACDDE0/2020\|EPI_ISL_630214\|20 | ···················· | ···················· | ···················· | ···················· | ···················· | ···················· |
| Q | 2410 | hCoV-19/England/MILK-AC94B2/2020\|EPI_ISL_630215\|20 | ···················· | ···················· | ···················· | ···················· | ···T················ | ···················· |
| Q | 2411 | hCoV-19/England/MILK-AC72C9/2020\|EPI_ISL_630216\|20 | ···················· | ···················· | ···················· | ···················· | ···················· | ···················· |
| Q | 2412 | hCoV-19/England/MILK-ACDC98/2020\|EPI_ISL_630217\|20 | ···················· | ···················· | ···················· | ···················· | ···················· | ···················· |
| Q | 2413 | hCoV-19/England/MILK-ACE20E/2020\|EPI_ISL_630218\|20 | ···················· | ···················· | ···················· | ···················· | ···················· | ···················· |
| Q | 2414 | hCoV-19/England/MILK-AC7627/2020\|EPI_ISL_630220\|20 | ···················· | ···················· | ···················· | ···················· | ···················· | ···················· |
| Q | 2415 | hCoV-19/England/MILK-AC74F0/2020\|EPI_ISL_630221\|20 | ···················· | ···················· | ···················· | ···················· | ···················· | ···················· |
| Q | 2416 | hCoV-19/England/MILK-ACCFAF/2020\|EPI_ISL_630222\|20 | ···················· | ···················· | ···················· | ···················· | ···················· | ···················· |
| Q | 2417 | hCoV-19/England/MILK-ACD8D0/2020\|EPI_ISL_630223\|20 | ···················· | ···················· | ···················· | ···················· | ···················· | ···················· |
| Q | 2418 | hCoV-19/England/MILK-ACD120/2020\|EPI_ISL_630225\|20 | ···················· | ···················· | ···················· | ···················· | ···················· | ···················· |
| Q | 2419 | hCoV-19/England/MILK-AC9458/2020\|EPI_ISL_630226\|20 | ···················· | ···················· | ···················· | ···················· | ···················· | ···················· |
| Q | 2420 | hCoV-19/England/MILK-ACDE92/2020\|EPI_ISL_630227\|20 | ···················· | ···················· | ···················· | ···················· | ···················· | ···················· |
| Q | 2421 | hCoV-19/England/MILK-ACDD1D/2020\|EPI_ISL_630228\|20 | ···················· | ···················· | ···················· | ···················· | ···················· | ···················· |
| Q | 2422 | hCoV-19/England/MILK-ACD09C/2020\|EPI_ISL_630229\|20 | ···················· | ···················· | ···················· | ···················· | ···················· | ···················· |
| Q | 2423 | hCoV-19/England/MILK-ACCE48/2020\|EPI_ISL_630230\|20 | ···················· | ···················· | ···················· | ···················· | ···················· | ···················· |
| Q | 2424 | hCoV-19/England/MILK-ACD17B/2020\|EPI_ISL_630231\|20 | ···················· | ···················· | ···················· | ···················· | ···················· | ···················· |
| Q | 2425 | hCoV-19/England/MILK-ACE0E6/2020\|EPI_ISL_630233\|20 | ···················· | ···················· | ···················· | ···················· | ·····T·············· | ···················· |
| Q | 2426 | hCoV-19/England/MILK-AC94A3/2020\|EPI_ISL_630234\|20 | ···················· | ···················· | ···················· | ···················· | ···················· | ···················· |
| Q | 2427 | hCoV-19/England/MILK-AC77F7/2020\|EPI_ISL_630258\|20 | ···················· | ···················· | ···················· | ···················· | ···················· | ···················· |
| Q | 2428 | hCoV-19/England/MILK-AC9485/2020\|EPI_ISL_630259\|20 | ···················· | ···················· | ···················· | ···················· | ···················· | ···················· |
| Q | 2429 | hCoV-19/England/MILK-ACD296/2020\|EPI_ISL_630260\|20 | ···················· | ···················· | ···················· | ···················· | ···················· | ···················· |
| Q | 2430 | hCoV-19/England/MILK-ACD8EF/2020\|EPI_ISL_630262\|20 | ···················· | ···················· | ···················· | ···················· | ···················· | ···················· |
| Q | 2431 | hCoV-19/England/MILK-AC9A92/2020\|EPI_ISL_630263\|20 | ···················· | ···················· | ···················· | ···················· | ···················· | ···················· |
| Q | 2432 | hCoV-19/England/MILK-ACCDA5/2020\|EPI_ISL_630264\|20 | ···················· | ···················· | ···················· | ···················· | ···················· | ···················· |
| Q | 2433 | hCoV-19/England/MILK-ACDA34/2020\|EPI_ISL_630265\|20 | ···················· | ···················· | ···················· | ···················· | ···················· | ···················· |
| Q | 2434 | hCoV-19/England/MILK-ACDAAD/2020\|EPI_ISL_630267\|20 | ···················· | ···················· | ···················· | ···················· | ···················· | ···················· |
| Q | 2435 | hCoV-19/England/MILK-ACE189/2020\|EPI_ISL_630268\|20 | ···················· | ···················· | ···················· | ···················· | ·····T·············· | ···················· |
| Q | 2436 | hCoV-19/England/MILK-ACE0D7/2020\|EPI_ISL_630269\|20 | ···················· | ···················· | ···················· | ···················· | ···················· | ···················· |
| Q | 2437 | hCoV-19/England/MILK-AC9A83/2020\|EPI_ISL_630270\|20 | ···················· | ···················· | ···················· | ···················· | ···················· | ···················· |
| Q | 2438 | hCoV-19/England/MILK-AC934C/2020\|EPI_ISL_630271\|20 | ···················· | ···················· | ···················· | ···················· | ···················· | ···················· |
| Q | 2439 | hCoV-19/England/MILK-AC7751/2020\|EPI_ISL_630272\|20 | ···················· | ···················· | ···················· | ···················· | ···················· | ···················· |
| Q | 2440 | hCoV-19/England/MILK-AC77BB/2020\|EPI_ISL_630273\|20 | ···················· | ···················· | ···················· | ···················· | ···················· | ···················· |
| Q | 2441 | hCoV-19/England/MILK-ACDDB3/2020\|EPI_ISL_630274\|20 | ···················· | ···················· | ···················· | ···················· | ···················· | ···················· |
| Q | 2442 | hCoV-19/England/MILK-ACCE93/2020\|EPI_ISL_630275\|20 | ···················· | ···················· | ···················· | ···················· | ···················· | ···················· |
| Q | 2443 | hCoV-19/England/MILK-ACDC3E/2020\|EPI_ISL_630277\|20 | ···················· | ···················· | ···················· | ···················· | ···················· | ···················· |
| Q | 2444 | hCoV-19/England/MILK-AC7311/2020\|EPI_ISL_630278\|20 | ···················· | ···················· | ···················· | ···················· | ···················· | ···················· |
| Q | 2445 | hCoV-19/England/MILK-ACCC5D/2020\|EPI_ISL_630279\|20 | ···················· | ···················· | ···················· | ···················· | ···················· | ···················· |
| Q | 2446 | hCoV-19/England/MILK-ACE17A/2020\|EPI_ISL_630280\|20 | ···················· | ···················· | ···················· | ···················· | ···················· | ···················· |
| Q | 2447 | hCoV-19/England/MILK-AC90EB/2020\|EPI_ISL_630281\|20 | ···················· | ···················· | ···················· | ···················· | ···················· | ···················· |
| Q | 2448 | hCoV-19/England/MILK-AC9A74/2020\|EPI_ISL_630282\|20 | ···················· | ···················· | ···················· | ···················· | ···················· | ···················· |
| Q | 2449 | hCoV-19/England/MILK-ACD15D/2020\|EPI_ISL_630283\|20 | ···················· | ···················· | ···················· | ···················· | ···················· | ···················· |
| Q | 2450 | hCoV-19/England/MILK-ACDF35/2020\|EPI_ISL_630284\|20 | ···················· | ···················· | ···················· | ···················· | ···················· | ···················· |
| Q | 2451 | hCoV-19/England/MILK-ACD928/2020\|EPI_ISL_630285\|20 | ···················· | ···················· | ···················· | ···················· | ···················· | ···················· |
| Q | 2452 | hCoV-19/England/MILK-ACDF26/2020\|EPI_ISL_630286\|20 | ···················· | ···················· | ···················· | ···················· | ·····T·············· | ···················· |
| Q | 2453 | hCoV-19/England/MILK-AC7593/2020\|EPI_ISL_630288\|20 | ···················· | ···················· | ···················· | ···················· | ···················· | ···················· |
| Q | 2454 | hCoV-19/England/MILK-AC7645/2020\|EPI_ISL_630289\|20 | ···················· | ·A·················· | ···················· | ···················· | ···················· | ···A················ |
| Q | 2455 | hCoV-19/England/MILK-ACCC7B/2020\|EPI_ISL_630290\|20 | ···················· | ···················· | ···················· | ···················· | ···················· | ···················· |
| Q | 2456 | hCoV-19/England/MILK-ACDB22/2020\|EPI_ISL_630291\|20 | ···················· | ···················· | ···················· | ···················· | ···················· | ···················· |
| Q | 2457 | hCoV-19/England/MILK-AC76AF/2020\|EPI_ISL_630292\|20 | ···················· | ···················· | ···················· | ···················· | ···T················ | ···················· |
| Q | 2458 | hCoV-19/England/MILK-AC7760/2020\|EPI_ISL_630293\|20 | ···················· | ···················· | ···················· | ···················· | ···················· | ···················· |
| Q | 2459 | hCoV-19/England/MILK-ACDECF/2020\|EPI_ISL_630294\|20 | ···················· | ···················· | ···················· | ···················· | ···················· | ···················· |
| Q | 2460 | hCoV-19/England/MILK-AC77AC/2020\|EPI_ISL_630295\|20 | ···················· | ···················· | ···················· | ···················· | ···················· | ···················· |
| Q | 2461 | hCoV-19/England/MILK-AC7672/2020\|EPI_ISL_630296\|20 | ···················· | ···················· | ···················· | ···················· | ·····T·············· | ···················· |
| Q | 2462 | hCoV-19/England/MILK-ACDB40/2020\|EPI_ISL_630297\|20 | ···················· | ···················· | ···················· | ···················· | ···················· | ···················· |
| Q | 2463 | hCoV-19/Wales/MILK-ACD0AB/2020\|EPI_ISL_630299\|2020 | ···················· | ···················· | ···················· | ···················· | ···················· | ···················· |
| Q | 2464 | hCoV-19/USA/NY-NYCPHL-000027/2020\|EPI_ISL_631523\|2 | ···················· | ···················· | ···················· | ···················· | ···················· | ···················· |
| Q | 2465 | hCoV-19/USA/NY-NYCPHL-000028/2020\|EPI_ISL_631524\|2 | ···················· | ······A············· | ···················· | ···················· | ···················· | ···················· |
| Q | 2466 | hCoV-19/USA/NY-NYCPHL-000029/2020\|EPI_ISL_631525\|2 | ···················· | ···················· | ···················· | ···················· | ···················· | ···················· |
| Q | 2467 | hCoV-19/USA/NY-NYCPHL-000030/2020\|EPI_ISL_631526\|2 | ···················· | ···················· | ···················· | ···················· | ···················· | ···················· |
| Q | 2468 | hCoV-19/USA/NY-NYCPHL-000031/2020\|EPI_ISL_631527\|2 | ···················· | ···················· | ···················· | ···················· | ···················· | ···················· |
| Q | 2469 | hCoV-19/USA/NY-NYCPHL-000032/2020\|EPI_ISL_631528\|2 | ···················· | ···················· | ···················· | ···················· | ···················· | ···················· |
| Q | 2470 | hCoV-19/USA/NY-NYCPHL-000033/2020\|EPI_ISL_631529\|2 | ···················· | ···················· | ···················· | ···················· | ···················· | ···················· |
| Q | 2471 | hCoV-19/USA/NY-NYCPHL-000034/2020\|EPI_ISL_631530\|2 | ···················· | ···················· | ···················· | ···················· | ···················· | ···················· |
| Q | 2472 | hCoV-19/USA/NY-NYCPHL-000036/2020\|EPI_ISL_631531\|2 | ···················· | ···················· | ···················· | ···················· | ···················· | ···················· |
| Q | 2473 | hCoV-19/USA/NY-NYCPHL-000037/2020\|EPI_ISL_631532\|2 | ···················· | ···················· | ···················· | ···················· | ···················· | ···················· |
| Q | 2474 | hCoV-19/USA/NY-NYCPHL-000038/2020\|EPI_ISL_631533\|2 | ···················· | ···················· | ···················· | ···················· | ···················· | ···················· |
| Q | 2475 | hCoV-19/USA/NY-NYCPHL-000039/2020\|EPI_ISL_631534\|2 | ···················· | ···················· | ···················· | ···················· | ···················· | ···················· |
| Q | 2476 | hCoV-19/USA/NY-NYCPHL-000041/2020\|EPI_ISL_631536\|2 | ···················· | ···················· | ···················· | ···················· | ···················· | ···················· |
| Q | 2477 | hCoV-19/USA/NY-NYCPHL-000042/2020\|EPI_ISL_631537\|2 | ···················· | ···················· | ···················· | ···················· | ···················· | ···················· |
| Q | 2478 | hCoV-19/USA/NY-NYCPHL-000044/2020\|EPI_ISL_631538\|2 | ···················· | ···················· | ···················· | ···················· | ···················· | ···················· |
| Q | 2479 | hCoV-19/USA/NY-NYCPHL-000045/2020\|EPI_ISL_631539\|2 | ···················· | ···················· | ···················· | ···················· | ···················· | ···················· |
| Q | 2480 | hCoV-19/USA/NY-NYCPHL-000046/2020\|EPI_ISL_631540\|2 | ···················· | ···················· | ···················· | ···················· | ···················· | ···················· |
| Q | 2481 | hCoV-19/USA/NY-NYCPHL-000047/2020\|EPI_ISL_631541\|2 | ···················· | ···················· | ···················· | ···················· | ···················· | ···················· |

Q 2482 hCoV-19/USA/NY-NYCPHL-000048/2020|EPI_ISL_631542|2 ···················· ···················· ···················· ···················· ···················· ····················
Q 2483 hCoV-19/USA/NY-NYCPHL-000049/2020|EPI_ISL_631543|2 ···················· ···················· ···················· ···················· ···················· ····················
Q 2484 hCoV-19/USA/NY-NYCPHL-000050/2020|EPI_ISL_631544|2 ···················· ···················· ···················· ···················· ···················· ····················
Q 2485 hCoV-19/USA/NY-NYCPHL-000051/2020|EPI_ISL_631545|2 ···················· ···················· ···················· ···················· ···················· ····················
Q 2486 hCoV-19/USA/NY-NYCPHL-000052/2020|EPI_ISL_631546|2 ···················· ···················· ···················· ···················· ···················· ····················
Q 2487 hCoV-19/USA/NY-NYCPHL-000053/2020|EPI_ISL_631547|2 ···················· ···················· ···················· ···················· ···················· ····················
Q 2488 hCoV-19/USA/NY-NYCPHL-000054/2020|EPI_ISL_631548|2 ···················· ···················· ···················· ···················· ···················· ····················
Q 2489 hCoV-19/USA/NY-NYCPHL-000055/2020|EPI_ISL_631549|2 ···················· ···················· ···················· ···················· ···················· ····················
Q 2490 hCoV-19/USA/NY-NYCPHL-000056/2020|EPI_ISL_631550|2 ···················· ···················· ···················· ···················· ···················· ····················
Q 2491 hCoV-19/USA/NY-NYCPHL-000057/2020|EPI_ISL_631551|2 ···················· ···················· ···················· ···················· ···················· ····················
Q 2492 hCoV-19/USA/NY-NYCPHL-000059/2020|EPI_ISL_631553|2 ···················· ···················· ···················· ···················· ···················· ····················
Q 2493 hCoV-19/USA/NY-NYCPHL-000060/2020|EPI_ISL_631554|2 ···················· ···················· ···················· ···················· ···················· ····················
Q 2494 hCoV-19/USA/NY-NYCPHL-000061/2020|EPI_ISL_631555|2 ···················· ···················· ···················· ···················· ···················· ····················
Q 2495 hCoV-19/USA/NY-NYCPHL-000062/2020|EPI_ISL_631556|2 ···················· ···················· ···················· ···················· ···················· ····················
Q 2496 hCoV-19/USA/NY-NYCPHL-000063/2020|EPI_ISL_631557|2 ···················· ···················· ···················· ···················· ···················· ····················
Q 2497 hCoV-19/USA/NY-NYCPHL-000065/2020|EPI_ISL_631558|2 ···················· ···················· ···················· ···················· ···················· ····················
Q 2498 hCoV-19/USA/NY-NYCPHL-000066/2020|EPI_ISL_631559|2 ···················· ···················· ···················· ···················· ···················· ····················
Q 2499 hCoV-19/USA/NY-NYCPHL-000067/2020|EPI_ISL_631560|2 ···················· ···················· ···················· ···················· ···················· ····················
Q 2500 hCoV-19/USA/NY-NYCPHL-000068/2020|EPI_ISL_631561|2 ···················· ···················· ···················· ···················· ···················· ····················
Q 2501 hCoV-19/USA/NY-NYCPHL-000069/2020|EPI_ISL_631562|2 ···················· ···················· ···················· ···················· ···················· ····················
Q 2502 hCoV-19/USA/NY-NYCPHL-000070/2020|EPI_ISL_631563|2 ···················· ···················· ···················· ···················· ···················· ····················
Q 2503 hCoV-19/USA/NY-NYCPHL-000071/2020|EPI_ISL_631564|2 ···················· ···················· ···················· ···················· ···················· ····················
Q 2504 hCoV-19/USA/NY-NYCPHL-000072/2020|EPI_ISL_631565|2 ···················· ···················· ···················· ···················· ···················· ····················
Q 2505 hCoV-19/USA/NY-NYCPHL-000073/2020|EPI_ISL_631566|2 ···················· ···················· ···················· ···················· ···················· ····················
Q 2506 hCoV-19/USA/NY-NYCPHL-000098/2020|EPI_ISL_631590|2 ···················· ···················· ···················· ···················· ···················· ····················
Q 2507 hCoV-19/USA/NY-NYCPHL-000099/2020|EPI_ISL_631591|2 ···················· ···················· ···················· ···················· ···················· ····················
Q 2508 hCoV-19/USA/NY-NYCPHL-000101/2020|EPI_ISL_631593|2 ···················· ···················· ···················· ···················· ···················· ····················
Q 2509 hCoV-19/USA/NY-NYCPHL-000102/2020|EPI_ISL_631594|2 ···················· ···················· ···················· ···················· ···················· ····················
Q 2510 hCoV-19/USA/NY-NYCPHL-000103/2020|EPI_ISL_631595|2 ···················· ···················· ···················· ···················· ···················· ····················
Q 2511 hCoV-19/USA/NY-NYCPHL-000104/2020|EPI_ISL_631596|2 ···················· ···················· ···················· ···················· ···················· ····················
Q 2512 hCoV-19/USA/NY-NYCPHL-000105/2020|EPI_ISL_631597|2 ···················· ···················· ···················· ···················· ···················· ····················
Q 2513 hCoV-19/USA/NY-NYCPHL-000106/2020|EPI_ISL_631598|2 ···················· ···················· ···················· ···················· ···················· ····················
Q 2514 hCoV-19/USA/NY-NYCPHL-000107/2020|EPI_ISL_631599|2 ···················· ···················· ···················· ···················· ···················· ····················
Q 2515 hCoV-19/USA/ID-UNM-IBL063/2020|EPI_ISL_632856|2020 ···················· ···················· ···················· ···················· ···················· ····················
Q 2516 hCoV-19/USA/ID-UNM-IBL021/2020|EPI_ISL_632857|2020 ···················· ···················· ···················· ···················· ···················· ····················
Q 2517 hCoV-19/USA/ID-UNM-IBL025/2020|EPI_ISL_632858|2020 ···················· ···················· ···················· ···················· ···················· ····················
Q 2518 hCoV-19/USA/ID-UNM-IBL022/2020|EPI_ISL_632859|2020 ···················· ···················· ···················· ···················· ···················· ····················
Q 2519 hCoV-19/USA/ID-UNM-IBL311/2020|EPI_ISL_632861|2020 ···················· ···················· ···················· ···················· ···················· ····················
Q 2520 hCoV-19/USA/ID-UNM-IBL068/2020|EPI_ISL_632863|2020 ···················· ···················· ···················· ···················· ···················· ····················
Q 2521 hCoV-19/USA/ID-UNM-IBL052/2020|EPI_ISL_632864|2020 ···················· ···················· ···················· ···················· ···················· ····················
Q 2522 hCoV-19/USA/ID-UNM-IBL041/2020|EPI_ISL_632865|2020 ···················· ·A·················· ···················· ···················· ···················· ····················
Q 2523 hCoV-19/USA/ID-UNM-IBL023/2020|EPI_ISL_632868|2020 ···················· ···················· ···················· ···················· ···················· ····················
Q 2524 hCoV-19/USA/ID-UNM-IBL026/2020|EPI_ISL_632870|2020 ···················· ···················· ···················· ···················· ···················· ····················
Q 2525 hCoV-19/USA/ID-UNM-IBL028/2020|EPI_ISL_632871|2020 ···················· ···················· ···················· ···················· ···················· ····················
Q 2526 hCoV-19/USA/ID-UNM-IBL273/2020|EPI_ISL_632872|2020 ···················· ···················· ···················· ···················· ···················· ····················
Q 2527 hCoV-19/USA/ID-UNM-IBL037/2020|EPI_ISL_632873|2020 ···················· ···················· ···················· ···················· ···················· ····················
Q 2528 hCoV-19/USA/ID-UNM-IBL038/2020|EPI_ISL_632876|2020 ···················· ···················· ···················· ···················· ···················· ····················
Q 2529 hCoV-19/USA/ID-UNM-IBL154/2020|EPI_ISL_632878|2020 ···················· ···················· ···················· ···················· ···················· ····················
Q 2530 hCoV-19/USA/ID-UNM-IBL150/2020|EPI_ISL_632879|2020 ···················· ···················· ···················· ···················· ···················· ····················
Q 2531 hCoV-19/USA/ID-UNM-IBL067/2020|EPI_ISL_632880|2020 ···················· ···················· ···················· ···················· ···················· ····················
Q 2532 hCoV-19/USA/ID-UNM-IBL039/2020|EPI_ISL_632881|2020 ···················· ···················· ···················· ···················· ···················· ····················
Q 2533 hCoV-19/USA/ID-UNM-IBL040/2020|EPI_ISL_632882|2020 ···················· ···················· ···················· ···················· ···················· ····················
Q 2534 hCoV-19/USA/ID-UNM-IBL061/2020|EPI_ISL_632883|2020 ···················· ···················· ···················· ···················· ···················· ····················
Q 2535 hCoV-19/USA/ID-UNM-IBL056/2020|EPI_ISL_632885|2020 ···················· ···················· ···················· ···················· ···················· ····················
Q 2536 hCoV-19/USA/ID-UNM-IBL062/2020|EPI_ISL_632886|2020 ···················· ···················· ···················· ···················· ···················· ····················
Q 2537 hCoV-19/USA/WI-UW-1611/2020|EPI_ISL_632891|2020-09 ···················· ···················· ···················· ···················· ···················· ····················
Q 2538 hCoV-19/USA/WI-UW-1613/2020|EPI_ISL_632892|2020-09 ···················· ···················· ···················· ···················· ···················· ····················
Q 2539 hCoV-19/USA/WI-UW-1614/2020|EPI_ISL_632893|2020-09 ···················· ···················· ···················· ···················· ···················· ····················
Q 2540 hCoV-19/USA/WI-UW-1768/2020|EPI_ISL_632895|2020-09 ···················· ···················· ···················· ···················· ···················· ····················
Q 2541 hCoV-19/USA/WI-UW-1877/2020|EPI_ISL_632896|2020-09 ···················· ···················· ···················· ···················· ···················· ····················
Q 2543 hCoV-19/Jordan/AM-ALSR-4040/2020|EPI_ISL_636400|20 ···················· ·A·················· ···················· ···················· ···················· ··A·················
Q 2544 hCoV-19/Jordan/AM-ALSR-4041/2020|EPI_ISL_636401|20 ···················· ···················· ···················· ···················· ···················· ····················
Q 2545 hCoV-19/Jordan/AM-ALSR-4042/2020|EPI_ISL_636402|20 ···················· ···················· ···················· ···················· ···················· ····················
Q 2546 hCoV-19/Jordan/AM-ALSR-4043/2020|EPI_ISL_636403|20 ···················· ···················· ···················· ···················· ···················· ····················
Q 2547 hCoV-19/Jordan/AM-ALSR-4044/2020|EPI_ISL_636404|20 ···················· ···················· ···················· ···················· ···················· ····················
Q 2548 hCoV-19/Jordan/AM-ALSR-4045/2020|EPI_ISL_636405|20 ···················· ···················· ···················· ···················· ···················· ····················
Q 2549 hCoV-19/Jordan/AM-ALSR-4046/2020|EPI_ISL_636406|20 ···················· ···················· ···················· ···················· ···················· ····················
Q 2550 hCoV-19/Jordan/AM-ALSR-4047/2020|EPI_ISL_636407|20 ···················· ···················· ···················· ···················· ···················· ····················
Q 2551 hCoV-19/Jordan/AM-ALSR-4048/2020|EPI_ISL_636408|20 ···················· ···················· ···················· ···················· ···················· ····················
Q 2552 hCoV-19/Jordan/AM-ALSR-4049/2020|EPI_ISL_636409|20 ···················· ···················· ···················· ···················· ···················· ····················
Q 2553 hCoV-19/Jordan/AM-ALSR-4050/2020|EPI_ISL_636410|20 ···················· ·A·················· ···················· ···················· ···················· ··A·················
Q 2554 hCoV-19/Jordan/AM-ALSR-4051/2020|EPI_ISL_636411|20 ···················· ·A·················· ···················· ···················· ···················· ··A·················
Q 2555 hCoV-19/Jordan/AM-ALSR-4052/2020|EPI_ISL_636412|20 ···················· ···················· ···················· ···················· ···················· ····················
Q 2556 hCoV-19/Jordan/AM-ALSR-4053/2020|EPI_ISL_636413|20 ···················· ···················· ···················· ···················· ···················· ····················
Q 2557 hCoV-19/Jordan/AM-ALSR-4054/2020|EPI_ISL_636414|20 ···················· ···················· ···················· ···················· ·····T·············· ····················
Q 2558 hCoV-19/Jordan/AM-ALSR-4055/2020|EPI_ISL_636415|20 ···················· ···················· ···················· ···················· ·····T·············· ····················
Q 2559 hCoV-19/Jordan/AM-ALSR-4056/2020|EPI_ISL_636416|20 ···················· ···················· ···················· ···················· ···················· ····················
Q 2560 hCoV-19/Jordan/AM-ALSR-4057/2020|EPI_ISL_636417|20 ···················· ···················· ···················· ···················· ···················· ····················
Q 2561 hCoV-19/Scotland/CVR5209/2020|EPI_ISL_627390|2020- ···················· ···················· ···················· ···NN···········N··· ···················· ···················· 1
Q 2562 hCoV-19/Scotland/CVR5214/2020|EPI_ISL_627391|2020- ···················· ···················· ···················· ···················· ···················· ····················

| | | | | | | | | |
|---|---|---|---|---|---|---|---|---|
| Q | 2563 hCoV-19/Scotland/CVR5217/2020\|EPI_ISL_627392\|2020- | ···················· | ···················· | ···················· | ···················· | ···················· | ···················· | |
| Q | 2564 hCoV-19/Scotland/CVR5219/2020\|EPI_ISL_627393\|2020- | ···················· | ···················· | ···················· | ···················· | ···················· | ···················· | |
| Q | 2565 hCoV-19/Scotland/CVR5220/2020\|EPI_ISL_627394\|2020- | ···················· | ···················· | ···················· | ···················· | ···················· | ···················· | |
| Q | 2566 hCoV-19/Scotland/CVR5223/2020\|EPI_ISL_627395\|2020- | ···················· | ···················· | ···················· | ···················· | ···················· | ···················· | |
| Q | 2567 hCoV-19/Scotland/CVR5224/2020\|EPI_ISL_627396\|2020- | ···················· | ···················· | ···················· | ···················· | ···················· | ···················· | |
| Q | 2569 hCoV-19/Scotland/CVR5226/2020\|EPI_ISL_627398\|2020- | ···················· | ···················· | ···················· | ···················· | ···················· | ···················· | |
| Q | 2571 hCoV-19/Scotland/CVR5248/2020\|EPI_ISL_627410\|2020- | ···················· | ···················· | ···················· | ···················· | ···················· | ···················· | |
| Q | 2572 hCoV-19/Scotland/CVR5249/2020\|EPI_ISL_627411\|2020- | ···················· | ···················· | ···················· | ···················· | ···················· | ···················· | |
| Q | 2573 hCoV-19/Scotland/CVR5254/2020\|EPI_ISL_627416\|2020- | ···················· | ···················· | ···················· | ···················· | ···················· | ···················· | |
| Q | 2574 hCoV-19/Scotland/CVR5255/2020\|EPI_ISL_627417\|2020- | ···················· | ···················· | ···················· | ···················· | T··················· | ···················· | |
| Q | 2575 hCoV-19/Scotland/CVR5259/2020\|EPI_ISL_627419\|2020- | ···················· | ···················· | ···················· | ···················· | ···················· | ···················· | |
| Q | 2577 hCoV-19/England/EXET-13896D/2020\|EPI_ISL_627431\|20 | ···················· | ····A··············· | ···················· | ···················· | ···················· | ···················· | |
| Q | 2578 hCoV-19/England/EXET-138A3D/2020\|EPI_ISL_627432\|20 | ···················· | ····A··············· | ···················· | ···················· | ···················· | ···················· | |
| Q | 2579 hCoV-19/England/EXET-138A6A/2020\|EPI_ISL_627433\|20 | ···················· | ···················· | ···················· | ···················· | ···················· | ···················· | |
| Q | 2580 hCoV-19/England/EXET-138AF1/2020\|EPI_ISL_627434\|20 | ···················· | ···················· | ···················· | ···················· | ···················· | ···················· | |
| Q | 2581 hCoV-19/England/EXET-138B85/2020\|EPI_ISL_627435\|20 | ···················· | ···················· | ···················· | ···················· | ···················· | ···················· | |
| Q | 2582 hCoV-19/England/EXET-138BA3/2020\|EPI_ISL_627436\|20 | ···················· | ···················· | ···················· | ···················· | ···················· | ···················· | |
| Q | 2583 hCoV-19/England/EXET-138C28/2020\|EPI_ISL_627437\|20 | ···················· | ···················· | ···················· | ···················· | ···················· | ···················· | |
| Q | 2584 hCoV-19/England/EXET-138C37/2020\|EPI_ISL_627438\|20 | ···················· | ···················· | ···················· | ···················· | ···················· | ···················· | |
| Q | 2585 hCoV-19/England/EXET-138C46/2020\|EPI_ISL_627439\|20 | ···················· | ···················· | ···················· | ···················· | ···················· | ···················· | |
| Q | 2586 hCoV-19/England/EXET-138C64/2020\|EPI_ISL_627440\|20 | ···················· | ···················· | ···················· | ···················· | ···················· | ···················· | |
| Q | 2587 hCoV-19/England/EXET-138C73/2020\|EPI_ISL_627441\|20 | ···················· | ···················· | ···················· | ···················· | ···················· | ···················· | |
| Q | 2588 hCoV-19/England/EXET-138D16/2020\|EPI_ISL_627442\|20 | ···················· | ···················· | ···················· | ···················· | ···················· | ···················· | |
| Q | 2589 hCoV-19/England/EXET-138D9E/2020\|EPI_ISL_627443\|20 | ···················· | ···················· | ···················· | ···················· | ···················· | ···················· | |
| Q | 2591 hCoV-19/England/LIVE-1E013F/2020\|EPI_ISL_627445\|20 | ···················· | ···················· | ···················· | ···················· | ···················· | ···················· | |
| Q | 2592 hCoV-19/Scotland/LIVE-1E02D2/2020\|EPI_ISL_627446\|2 | ···················· | ···················· | ···················· | ···················· | ···················· | ···················· | |
| Q | 2593 hCoV-19/England/LIVE-1E4326/2020\|EPI_ISL_627447\|20 | ···················· | ···················· | ···················· | ···················· | ···················· | ···················· | |
| Q | 2594 hCoV-19/England/LIVE-1E45A8/2020\|EPI_ISL_627448\|20 | ···················· | ···················· | ···················· | ···················· | ···················· | ···················· | |
| Q | 2595 hCoV-19/England/LIVE-DA4B1F/2020\|EPI_ISL_627449\|20 | ···················· | ···················· | ···················· | ···················· | ···················· | ···················· | |
| Q | 2596 hCoV-19/England/LOND-1262C21/2020\|EPI_ISL_627450\|2 | ···················· | ···················· | ···················· | ···················· | ·····T·············· | ···················· | |
| Q | 2597 hCoV-19/England/LOND-1262C6D/2020\|EPI_ISL_627451\|2 | ···················· | ···················· | ···················· | ···················· | ···················· | ···················· | |
| Q | 2598 hCoV-19/England/LOND-1262C8B/2020\|EPI_ISL_627452\|2 | ···················· | ···················· | ···················· | ···················· | ···················· | ···················· | |
| Q | 2599 hCoV-19/England/LOND-1262CA9/2020\|EPI_ISL_627453\|2 | ···················· | ···················· | ···················· | ···················· | ···················· | ···················· | |
| Q | 2600 hCoV-19/England/LOND-1262D3D/2020\|EPI_ISL_627454\|2 | ···················· | ···················· | ···················· | ···················· | ···················· | ···················· | |
| Q | 2601 hCoV-19/England/LOND-1262D5B/2020\|EPI_ISL_627455\|2 | ···················· | ···················· | ···················· | ···················· | ···················· | ···················· | |
| Q | 2602 hCoV-19/England/LOND-1262D79/2020\|EPI_ISL_627456\|2 | ···················· | ···················· | ···················· | ···················· | ···················· | ···················· | |
| Q | 2603 hCoV-19/England/LOND-1262D88/2020\|EPI_ISL_627457\|2 | ···················· | ···················· | ···················· | ···················· | ···················· | ···················· | |
| Q | 2604 hCoV-19/England/LOND-1262D97/2020\|EPI_ISL_627458\|2 | ···················· | ···················· | ···················· | ···················· | ·····T·············· | ···············A···· | |
| Q | 2605 hCoV-19/England/LOND-1262DB5/2020\|EPI_ISL_627459\|2 | ···················· | ···················· | ···················· | ···················· | ···················· | ···················· | |
| Q | 2606 hCoV-19/England/LOND-1262DE2/2020\|EPI_ISL_627460\|2 | ···················· | ·A·················· | ···················· | ···················· | ···················· | ··A················· | |
| Q | 2607 hCoV-19/England/LOND-1262E0D/2020\|EPI_ISL_627461\|2 | ···················· | ·A·················· | ···················· | ···················· | ···················· | ···················· | |
| Q | 2608 hCoV-19/England/LOND-1262E49/2020\|EPI_ISL_627462\|2 | ···················· | ···················· | ···················· | ···················· | ···················· | ···················· | |
| Q | 2609 hCoV-19/England/LOND-1262EA3/2020\|EPI_ISL_627463\|2 | ···················· | ···················· | ···················· | ···················· | ···················· | ···················· | |
| Q | 2610 hCoV-19/England/LOND-126C9A/2020\|EPI_ISL_627464\|20 | ···················· | ···················· | ···················· | ···················· | ···················· | ···················· | |
| Q | 2611 hCoV-19/England/LOND-DC2F8/2020\|EPI_ISL_627465\|202 | ···················· | ·A·················· | ···················· | ···················· | ···················· | ··A················· | |
| Q | 2612 hCoV-19/England/LOND-DC304/2020\|EPI_ISL_627466\|202 | ···················· | ·A·················· | ···················· | ···················· | ···················· | ··A················· | |
| Q | 2613 hCoV-19/England/LOND-DC322/2020\|EPI_ISL_627467\|202 | ···················· | ·A·················· | ···················· | ···················· | ···················· | ···················· | |
| Q | 2614 hCoV-19/England/LOND-DC331/2020\|EPI_ISL_627468\|202 | ···················· | ·A·················· | ···················· | ···················· | ···················· | ···················· | |
| Q | 2615 hCoV-19/England/LOND-DC36E/2020\|EPI_ISL_627469\|202 | ···················· | ·A·················· | ···················· | ···················· | ···················· | ···················· | |
| Q | 2616 hCoV-19/England/LOND-DC37D/2020\|EPI_ISL_627470\|202 | ···················· | ···················· | ···················· | ···················· | ···················· | ···················· | |
| Q | 2617 hCoV-19/England/LOND-DC39B/2020\|EPI_ISL_627471\|202 | ···················· | ···················· | ···················· | ···················· | ···················· | ···················· | |
| Q | 2618 hCoV-19/England/MILK-A18A1C/2020\|EPI_ISL_627472\|20 | ···················· | ···················· | ···················· | ···················· | ···················· | ···················· | |
| Q | 2619 hCoV-19/England/MILK-A18A2B/2020\|EPI_ISL_627473\|20 | ···················· | ···················· | ···················· | ···················· | ···················· | ···················· | |
| Q | 2620 hCoV-19/England/MILK-A18A49/2020\|EPI_ISL_627474\|20 | ···················· | ···················· | ···················· | ···················· | ···················· | ···················· | |
| Q | 2621 hCoV-19/England/MILK-A18A67/2020\|EPI_ISL_627475\|20 | ···················· | ···················· | ···················· | ···················· | ···················· | ···················· | |
| Q | 2622 hCoV-19/England/MILK-A18A76/2020\|EPI_ISL_627476\|20 | ···················· | ···················· | ···················· | ···················· | ···················· | ···················· | |
| Q | 2623 hCoV-19/England/MILK-A18A94/2020\|EPI_ISL_627477\|20 | ···················· | ···················· | ···················· | ···················· | ···················· | ···················· | |
| Q | 2624 hCoV-19/England/MILK-A18AD0/2020\|EPI_ISL_627478\|20 | ···················· | ···················· | ···················· | ···················· | ···················· | ···················· | |
| Q | 2625 hCoV-19/England/MILK-A18C8F/2020\|EPI_ISL_627479\|20 | ···················· | ···················· | ···················· | ···················· | ···················· | ···················· | |
| Q | 2626 hCoV-19/England/MILK-A18CDA/2020\|EPI_ISL_627480\|20 | ···················· | ···················· | ···················· | ···················· | ···················· | ···················· | |
| Q | 2627 hCoV-19/England/MILK-A18CE9/2020\|EPI_ISL_627481\|20 | ···················· | ···················· | ···················· | ···················· | ···················· | ···················· | |
| Q | 2628 hCoV-19/England/MILK-A18E98/2020\|EPI_ISL_627482\|20 | ···················· | ···················· | ···················· | ···················· | ···················· | ···················· | |
| Q | 2629 hCoV-19/England/MILK-A18EB6/2020\|EPI_ISL_627483\|20 | ···················· | ·A·················· | ···················· | ···················· | ···················· | ··A················· | |
| Q | 2630 hCoV-19/England/MILK-A190BF/2020\|EPI_ISL_627484\|20 | ···················· | ···················· | ···················· | ···················· | ···················· | ···················· | |
| Q | 2631 hCoV-19/England/MILK-A190DD/2020\|EPI_ISL_627485\|20 | ···················· | ···················· | ···················· | ···················· | ···················· | ···················· | |
| Q | 2632 hCoV-19/England/MILK-A19116/2020\|EPI_ISL_627486\|20 | ···················· | ···················· | ···················· | ···················· | ···················· | NNN················· | 1 |
| Q | 2634 hCoV-19/England/MILK-A191E9/2020\|EPI_ISL_627488\|20 | ···················· | ·R·················· | ···················· | ···················· | ···················· | ··R················· | |
| Q | 2635 hCoV-19/England/MILK-A19565/2020\|EPI_ISL_627489\|20 | ···················· | ···················· | ···················· | ···················· | ···················· | ···················· | |
| Q | 2636 hCoV-19/Wales/MILK-A195A1/2020\|EPI_ISL_627490\|2020 | ···················· | ···················· | ···················· | ···················· | ···················· | ···················· | |
| Q | 2637 hCoV-19/England/MILK-A19644/2020\|EPI_ISL_627491\|20 | ···················· | ···················· | ···················· | ···················· | ···················· | ···················· | |
| Q | 2638 hCoV-19/England/MILK-A6A073/2020\|EPI_ISL_627492\|20 | ···················· | ···················· | ···················· | ···················· | ···················· | ···················· | |
| Q | 2639 hCoV-19/England/MILK-A6A0FB/2020\|EPI_ISL_627493\|20 | ···················· | ···················· | ···················· | ···················· | ···················· | ···················· | |
| Q | 2640 hCoV-19/England/MILK-A6A116/2020\|EPI_ISL_627494\|20 | ···················· | ···················· | ···················· | ···················· | ···················· | ···················· | |
| Q | 2641 hCoV-19/England/MILK-A6A222/2020\|EPI_ISL_627495\|20 | ···················· | ···················· | ···················· | ···················· | ···················· | ···················· | |
| Q | 2642 hCoV-19/England/MILK-A6A240/2020\|EPI_ISL_627496\|20 | ···················· | ···················· | ···················· | ···················· | ···················· | ···················· | |
| Q | 2643 hCoV-19/England/MILK-A6A2B9/2020\|EPI_ISL_627497\|20 | ···················· | ···················· | ···················· | ···················· | ···················· | ···················· | |
| Q | 2644 hCoV-19/England/MILK-A6A301/2020\|EPI_ISL_627498\|20 | ···················· | ···················· | ···················· | ···················· | ···················· | ···················· | |
| Q | 2645 hCoV-19/England/MILK-A6A32F/2020\|EPI_ISL_627499\|20 | ···················· | ···················· | ···················· | ···················· | ···················· | ···················· | |
| Q | 2646 hCoV-19/England/QEUH-A6446E/2020\|EPI_ISL_628700\|20 | ···················· | ···················· | ···················· | ···················· | ···················· | ···················· | |
| Q | 2647 hCoV-19/England/QEUH-AE4C5F/2020\|EPI_ISL_633228\|20 | ···················· | ···················· | ···················· | ···················· | ···················· | ···················· | |

```
Q    2648 hCoV-19/England/CAMC-AEA697/2020|EPI_ISL_633229|20    ....................  ....................  ....................  ....................  ....................  ....................
Q    2649 hCoV-19/England/QEUH-AE50F8/2020|EPI_ISL_633230|20    ....................  .A..................  ....................  ....................  ....................  ....................
Q    2650 hCoV-19/England/CAMC-AEA961/2020|EPI_ISL_633231|20    ....................  ....................  ....................  ....................  ....................  ....................
Q    2651 hCoV-19/England/QEUH-AE4F83/2020|EPI_ISL_633232|20    ....................  ....................  ....................  ....................  ....................  ....................
Q    2652 hCoV-19/England/QEUH-AE4D6B/2020|EPI_ISL_633233|20    ....................  ....................  ....................  ....................  ....................  ..A.................
Q    2653 hCoV-19/England/CAMC-AEA7B2/2020|EPI_ISL_633234|20    ....................  ....................  ....................  ....................  ....................  ....................
Q    2654 hCoV-19/England/CAMC-AEAC59/2020|EPI_ISL_633235|20    ....................  ....................  ....................  ....................  ....................  ....................
Q    2655 hCoV-19/England/CAMC-AEA6A6/2020|EPI_ISL_633236|20    ....................  ....................  ....................  ....................  ....................  ....................
Q    2656 hCoV-19/England/QEUH-AE5025/2020|EPI_ISL_633237|20    ....................  ....................  ....................  ....................  ....................  ....................
Q    2657 hCoV-19/England/CAMC-AEA73A/2020|EPI_ISL_633239|20    ....................  ....................  ....................  ....................  ....................  ....................
Q    2658 hCoV-19/England/QEUH-AE4C40/2020|EPI_ISL_633240|20    ....................  ....................  ....................  ....................  ....................  ....................
Q    2659 hCoV-19/England/QEUH-AE524D/2020|EPI_ISL_633241|20    ....................  ....................  ....................  ....................  ....................  ....................
Q    2660 hCoV-19/England/CAMC-AEAD0B/2020|EPI_ISL_633242|20    ....................  ....................  ....................  ....................  ....................  ....................
Q    2661 hCoV-19/England/QEUH-AE5210/2020|EPI_ISL_633243|20    ....................  ....................  ....................  ....................  ....................  ....................
Q    2662 hCoV-19/England/QEUH-AE5043/2020|EPI_ISL_633244|20    ....................  ....................  ....................  ....................  ....................  ....................
Q    2663 hCoV-19/England/CAMC-AEA916/2020|EPI_ISL_633245|20    ....................  ....................  ....................  ....................  ......T.............  ....................
Q    2664 hCoV-19/England/CAMC-AEA934/2020|EPI_ISL_633246|20    ....................  ....................  ....................  ....................  ......T.............  ....................
Q    2665 hCoV-19/England/CAMC-AEA828/2020|EPI_ISL_633247|20    ....................  ....................  ....................  ....................  ....................  ....................
Q    2666 hCoV-19/England/CAMC-AEA98F/2020|EPI_ISL_633248|20    ....................  ....................  ....................  ....................  ....................  ....................
Q    2667 hCoV-19/England/CAMC-AEAC2C/2020|EPI_ISL_633249|20    ....................  .A..................  ....................  ....................  ....................  ....................
Q    2668 hCoV-19/England/CAMC-AEA7A3/2020|EPI_ISL_633250|20    ....................  ....................  ....................  ....................  ....................  ....................
Q    2669 hCoV-19/England/QEUH-AE4BBC/2020|EPI_ISL_633251|20    ....................  ....................  ....................  ....................  ......T.............  ....................
Q    2670 hCoV-19/England/CAMC-AEA8CE/2020|EPI_ISL_633252|20    ....................  ....................  ....................  ....................  ......T.............  ....................
Q    2671 hCoV-19/England/CAMC-AEAD47/2020|EPI_ISL_633253|20    ....................  ....................  ....................  ....................  ....................  ....................
Q    2672 hCoV-19/England/CAMC-AEA9E9/2020|EPI_ISL_633254|20    ....................  ....................  ....................  ....................  ....................  ....................
Q    2673 hCoV-19/England/CAMC-AEAB3E/2020|EPI_ISL_633255|20    ....................  .A..................  ....................  ....................  ....................  ..A.................
Q    2674 hCoV-19/England/QEUH-AE4ED1/2020|EPI_ISL_633256|20    ....................  ....................  ....................  ....................  ....................  ....................
Q    2675 hCoV-19/England/CAMC-AEB0D4/2020|EPI_ISL_633257|20    ....................  ....................  ....................  ....................  ....................  ....................
Q    2676 hCoV-19/England/CAMC-AEB06B/2020|EPI_ISL_633258|20    ....................  ....................  ....................  ....................  ....................  ....................
Q    2677 hCoV-19/England/CAMC-AEA9DA/2020|EPI_ISL_633260|20    ....................  ....................  ....................  ....................  ....................  ....................
Q    2678 hCoV-19/England/QEUH-AE4A82/2020|EPI_ISL_633261|20    ....................  ....................  ....................  ....................  ......T.............  ....................
Q    2679 hCoV-19/England/CAMC-AEA6E2/2020|EPI_ISL_633263|20    ....................  ....................  ....................  ....................  ....................  ....................
Q    2680 hCoV-19/England/CAMC-AEAD65/2020|EPI_ISL_633264|20    ....................  ....................  ....................  ....................  ....................  ....................
Q    2681 hCoV-19/England/CAMC-AEA785/2020|EPI_ISL_633265|20    ....................  ....................  ....................  ....................  ....................  ....................
Q    2682 hCoV-19/England/CAMC-AEA776/2020|EPI_ISL_633266|20    ....................  ....................  ....................  ....................  ....................  ....................
Q    2683 hCoV-19/England/CAMC-AEACE0/2020|EPI_ISL_633267|20    ....................  ....................  ....................  ....................  ....................  ....................
Q    2684 hCoV-19/England/QEUH-AE4BF8/2020|EPI_ISL_633268|20    ....................  ....................  ....................  ....................  ....................  ....................
Q    2685 hCoV-19/England/QEUH-AE5289/2020|EPI_ISL_633269|20    ....................  ....A...............  ....................  ....................  ....................  ....................
Q    2686 hCoV-19/England/CAMC-AEADFC/2020|EPI_ISL_633270|20    ....................  ....................  ....................  ....................  ....................  ....................
Q    2687 hCoV-19/England/QEUH-AE4CAA/2020|EPI_ISL_633271|20    ....................  ....................  ....................  ....................  ....................  ....................
Q    2688 hCoV-19/England/QEUH-AE4C9B/2020|EPI_ISL_633272|20    ....................  ....................  ....................  ....................  ....................  ....................
Q    2689 hCoV-19/England/CAMC-AEAD38/2020|EPI_ISL_633273|20    ....................  ....................  ....................  ....................  ....................  ....................
Q    2690 hCoV-19/England/QEUH-AE4D10/2020|EPI_ISL_633274|20    ....................  ....................  ....................  ....................  ....................  ....................
Q    2691 hCoV-19/England/CAMC-AEA8A0/2020|EPI_ISL_633275|20    ....................  ....................  ....................  ....................  ......T.............  ....................
Q    2692 hCoV-19/England/QEUH-AE4D3E/2020|EPI_ISL_633276|20    ....................  ....................  ....................  ....................  ....................  ....................
Q    2693 hCoV-19/England/CAMC-AEAA22/2020|EPI_ISL_633277|20    ....................  ....................  ....................  ....................  ....................  ....................
Q    2694 hCoV-19/England/CAMC-AEA6B5/2020|EPI_ISL_633278|20    ....................  ....................  ....................  ....................  ....................  ....................
Q    2695 hCoV-19/England/QEUH-AE4EFF/2020|EPI_ISL_633279|20    ....................  ....................  ....................  ....................  ....................  ....................
Q    2696 hCoV-19/England/QEUH-AE5070/2020|EPI_ISL_633280|20    ....................  ....................  ....................  ....................  ....................  ....................
Q    2697 hCoV-19/England/CAMC-AEA7EF/2020|EPI_ISL_633281|20    ....................  ....................  ....................  ....................  ....................  ....................
Q    2698 hCoV-19/England/QEUH-AE5052/2020|EPI_ISL_633283|20    ....................  ....................  ....................  ....................  ....................  ....................
Q    2699 hCoV-19/England/CAMC-AEA6F1/2020|EPI_ISL_633284|20    ....................  ....................  ....................  ....................  ....................  ....................
Q    2700 hCoV-19/England/QEUH-AE50CB/2020|EPI_ISL_633285|20    ....................  ....................  ....................  ....................  ....................  ....................
Q    2701 hCoV-19/England/CAMC-AEAA8C/2020|EPI_ISL_633286|20    ....................  ....................  ....................  ....................  ....................  ....................
Q    2702 hCoV-19/England/QEUH-AE49C1/2020|EPI_ISL_633287|20    ....................  ....................  ....................  ....................  ....................  ....................
Q    2703 hCoV-19/England/CAMC-AEAB89/2020|EPI_ISL_633288|20    ....................  ....................  ....................  ....................  ....................  ....................
Q    2704 hCoV-19/England/QEUH-AE4F0B/2020|EPI_ISL_633289|20    ....................  ....................  ....................  ....................  ....................  ....................
Q    2705 hCoV-19/England/CAMC-AEB04D/2020|EPI_ISL_633291|20    ....................  ....................  ....................  ....................  ....................  ....................
Q    2706 hCoV-19/England/CAMC-AEA62E/2020|EPI_ISL_633292|20    ....................  ....................  ....................  ....................  ....................  ....................
Q    2707 hCoV-19/England/QEUH-AE4E68/2020|EPI_ISL_633293|20    ....................  ....................  ....................  ....................  ....................  ....................
Q    2708 hCoV-19/England/CAMC-AEAC77/2020|EPI_ISL_633294|20    ....................  .A..................  ....................  ....................  ....................  ..A.................
Q    2709 hCoV-19/England/CAMC-AEAC0E/2020|EPI_ISL_633295|20    ....................  ....................  ....................  ....................  ....................  ....................
Q    2710 hCoV-19/England/QEUH-AE4DD4/2020|EPI_ISL_633296|20    ....................  ....................  ....................  ....................  ....................  ....................
Q    2711 hCoV-19/England/CAMC-AEA794/2020|EPI_ISL_633298|20    ....................  ....................  ....................  ....................  ....................  ....................
Q    2712 hCoV-19/England/CAMC-AEB20B/2020|EPI_ISL_633299|20    ....................  ....................  ....................  ....................  ....................  ....................
Q    2713 hCoV-19/England/MILK-AC7E89/2020|EPI_ISL_634500|20    ....................  ....................  ....................  ....................  ....................  ....................
Q    2714 hCoV-19/England/MILK-ACD3B1/2020|EPI_ISL_634501|20    ....................  ....................  ....................  ....................  ......T.............  ....................
Q    2715 hCoV-19/England/MILK-AC80DD/2020|EPI_ISL_634502|20    ....................  .A..................  ....................  ....................  ....................  ..A.................
Q    2716 hCoV-19/England/MILK-ACD779/2020|EPI_ISL_634503|20    ....................  ....................  ....................  ....................  ....................  ....................
Q    2717 hCoV-19/England/MILK-AC7BCE/2020|EPI_ISL_634504|20    ....................  ....................  ....................  ....................  ....................  ....................
Q    2718 hCoV-19/England/MILK-ACCBD8/2020|EPI_ISL_634505|20    ....................  ....................  ....................  ....................  ....................  ....................
Q    2719 hCoV-19/England/MILK-ACC4EC/2020|EPI_ISL_634506|20    ....................  ....................  ....................  ....................  ....................  ....................
Q    2720 hCoV-19/England/MILK-ACC525/2020|EPI_ISL_634507|20    ....................  ....................  ....................  ....................  ....................  ....................
Q    2721 hCoV-19/England/MILK-AD000F/2020|EPI_ISL_634508|20    ....................  .A..................  ....................  ....................  ....................  ..A.................
Q    2722 hCoV-19/England/MILK-ACD1C6/2020|EPI_ISL_634509|20    ....................  .A..................  ....................  ....................  ....................  ..A.................
Q    2723 hCoV-19/England/MILK-ACC6B9/2020|EPI_ISL_634511|20    ....................  ....................  ....................  ....................  ....................  ....................
Q    2724 hCoV-19/England/MILK-AC7C61/2020|EPI_ISL_634512|20    ....................  ....................  ....................  ....................  ......T.............  ....................
Q    2725 hCoV-19/England/MILK-AC7E98/2020|EPI_ISL_634513|20    ....................  .A..................  ....................  ....................  ....................  ..A.................
Q    2726 hCoV-19/England/MILK-ACD524/2020|EPI_ISL_634514|20    ....................  ....................  ....................  ....................  ....................  ....................
Q    2727 hCoV-19/England/QEUH-AD5A57/2020|EPI_ISL_634559|20    ....................  ....................  ....................  ....................  ....................  ....................
```

Q 2728 hCoV-19/England/QEUH-AD5F3A/2020|EPI_ISL_634560|20 ···················· ···················· ····················· ···················· ······T············· ····················
Q 2729 hCoV-19/England/QEUH-AD3FD2/2020|EPI_ISL_634561|20 ···················· ···················· ····················· ···················· ······T············· ····················
Q 2730 hCoV-19/England/QEUH-AD19C7/2020|EPI_ISL_634562|20 ···················· ···················· ····················· ···················· ···················· ····················
Q 2731 hCoV-19/England/QEUH-AD1086/2020|EPI_ISL_634563|20 ···················· ···················· ····················· ···················· ···················· ····················
Q 2732 hCoV-19/England/QEUH-AD1A6A/2020|EPI_ISL_634564|20 ···················· ···················· ····················· ···················· ···················· ····················
Q 2733 hCoV-19/England/QEUH-AD102C/2020|EPI_ISL_634565|20 ···················· ···················· ····················· ···················· ···················· ····················
Q 2734 hCoV-19/England/QEUH-AD4399/2020|EPI_ISL_634566|20 ···················· ···················· ····················· ···················· ···················· ····················
Q 2735 hCoV-19/England/QEUH-AD5ED3/2020|EPI_ISL_634567|20 ···················· ·A·················· ····················· ···················· ···················· ···A················
Q 2736 hCoV-19/England/QEUH-AD440F/2020|EPI_ISL_634568|20 ···················· ···················· ····················· ···················· ···················· ····················
Q 2737 hCoV-19/England/QEUH-AD4663/2020|EPI_ISL_634569|20 ···················· ···················· ····················· ···················· ······T············· ····················
Q 2738 hCoV-19/England/QEUH-AD5EA6/2020|EPI_ISL_634570|20 ···················· ···················· ····················· ···················· ···················· ····················
Q 2739 hCoV-19/England/QEUH-AD4C43/2020|EPI_ISL_634571|20 ···················· ···················· ····················· ···················· ···················· ····················
Q 2740 hCoV-19/England/QEUH-AD5CAC/2020|EPI_ISL_634572|20 ···················· ···················· ····················· ···················· ···················· ····················
Q 2741 hCoV-19/England/QEUH-AD5A39/2020|EPI_ISL_634573|20 ···················· ···················· ····················· ···················· ···················· ····················
Q 2742 hCoV-19/England/QEUH-AD1A79/2020|EPI_ISL_634574|20 ···················· ···················· ····················· ···················· ···················· ····················
Q 2743 hCoV-19/England/QEUH-AD1921/2020|EPI_ISL_634575|20 ···················· ···················· ····················· ···················· ···················· ····················
Q 2744 hCoV-19/England/QEUH-AD5CE8/2020|EPI_ISL_634576|20 ···················· ···················· ····················· ···················· ···················· ····················
Q 2745 hCoV-19/England/QEUH-AD18E8/2020|EPI_ISL_634578|20 ···················· ···················· ····················· ···················· ···················· ····················
Q 2746 hCoV-19/England/QEUH-AD41BD/2020|EPI_ISL_634579|20 ···················· ···················· ····················· ···················· ···················· ····················
Q 2747 hCoV-19/Scotland/QEUH-AD53B6/2020|EPI_ISL_634580|2 ···················· ···················· ····················· ···················· ···················· ····················
Q 2748 hCoV-19/England/QEUH-AD5E3D/2020|EPI_ISL_634581|20 ···················· ···················· ····················· ···················· ···················· ····················
Q 2749 hCoV-19/England/QEUH-AD4C61/2020|EPI_ISL_634582|20 ···················· ···················· ····················· ···················· ···················· ····················
Q 2750 hCoV-19/England/QEUH-AD151E/2020|EPI_ISL_634583|20 ···················· ···················· ····················· ···················· ···················· ····················
Q 2751 hCoV-19/England/QEUH-AD16B1/2020|EPI_ISL_634584|20 ···················· ···················· ····················· ···················· ···················· ····················
Q 2752 hCoV-19/England/QEUH-AD5E6A/2020|EPI_ISL_634585|20 ···················· ···················· ····················· ···················· ···················· ····················
Q 2753 hCoV-19/England/QEUH-AD1A88/2020|EPI_ISL_634586|20 ···················· ···················· ····················· ···················· ···················· ····················
Q 2754 hCoV-19/England/QEUH-AD5AFD/2020|EPI_ISL_634587|20 ···················· ···················· ····················· ···················· ······K············· ····················
Q 2755 hCoV-19/England/QEUH-AD1F01/2020|EPI_ISL_634588|20 ···················· ···················· ····················· ···················· ···················· ····················
Q 2756 hCoV-19/England/QEUH-AD43C6/2020|EPI_ISL_634589|20 ···················· ···················· ····················· ···················· ···················· ····················
Q 2757 hCoV-19/England/QEUH-AD1A2E/2020|EPI_ISL_634591|20 ···················· ···················· ····················· ···················· ···················· ····················
Q 2758 hCoV-19/England/QEUH-AD4BDD/2020|EPI_ISL_634592|20 ···················· ···················· ····················· ···················· ···················· ····················
Q 2759 hCoV-19/England/QEUH-AD5F49/2020|EPI_ISL_634593|20 ···················· ···················· ····················· ···················· ···················· ····················
Q 2760 hCoV-19/England/QEUH-AD58B7/2020|EPI_ISL_634594|20 ···················· ···················· ····················· ···················· ···················· ····················
Q 2761 hCoV-19/England/QEUH-AD0FF3/2020|EPI_ISL_634595|20 ···················· ···················· ····················· ···················· ···················· ····················
Q 2762 hCoV-19/England/QEUH-AD1A1F/2020|EPI_ISL_634596|20 ···················· ···················· ····················· ···················· ···················· ····················
Q 2763 hCoV-19/England/QEUH-AD5E4C/2020|EPI_ISL_634597|20 ···················· ···················· ····················· ···················· ···················· ····················
Q 2764 hCoV-19/England/QEUH-AD5AA2/2020|EPI_ISL_634598|20 ···················· ···················· ····················· ···················· ···················· ····················
Q 2765 hCoV-19/England/QEUH-AD0FC6/2020|EPI_ISL_634599|20 ···················· ···················· ····················· ···················· ····T··············· ····················
Q 2766 hCoV-19/USA/CA-ALSR-3032/2020|EPI_ISL_635801|2020- ···················· ···················· ····················· ···················· ···················· ····················
Q 2767 hCoV-19/USA/CA-ALSR-3033/2020|EPI_ISL_635802|2020- ···················· ·A·················· ····················· ···················· ···················· ····················
Q 2768 hCoV-19/USA/CA-ALSR-3034/2020|EPI_ISL_635803|2020- ···················· ·A·················· ····················· ···················· ···················· ····················
Q 2769 hCoV-19/USA/CA-ALSR-3035/2020|EPI_ISL_635804|2020- ···················· ·A·················· ····················· ···················· ···················· ····················
Q 2770 hCoV-19/USA/CA-ALSR-3038/2020|EPI_ISL_635807|2020- ···················· ·A·················· ····················· ···················· ···················· ····················
Q 2771 hCoV-19/USA/CA-ALSR-3040/2020|EPI_ISL_635808|2020- ···················· ·A·················· ····················· ···················· ···················· ····················
Q 2772 hCoV-19/USA/CA-ALSR-3043/2020|EPI_ISL_635809|2020- ···················· ···················· ····················· ···················· ···················· ····················
Q 2773 hCoV-19/USA/CA-ALSR-3045/2020|EPI_ISL_635810|2020- ···················· ·A·················· ····················· ···················· ···················· ····················
Q 2774 hCoV-19/USA/CA-ALSR-3046/2020|EPI_ISL_635811|2020- ···················· ···················· ····················· ···················· ···················· ····················
Q 2775 hCoV-19/USA/CA-ALSR-3047/2020|EPI_ISL_635812|2020- ···················· ·A·················· ····················· ···················· ···················· ····················
Q 2776 hCoV-19/USA/CA-ALSR-3048/2020|EPI_ISL_635813|2020- ···················· ···················· ····················· ···················· ···················· ····················
Q 2777 hCoV-19/USA/CA-ALSR-3050/2020|EPI_ISL_635814|2020- ···················· ···················· ····················· ···················· ···················· ····················
Q 2778 hCoV-19/USA/CA-ALSR-3051/2020|EPI_ISL_635815|2020- ···················· ·A·················· ····················· ···················· ···················· ····················
Q 2779 hCoV-19/USA/CA-ALSR-3052/2020|EPI_ISL_635816|2020- ···················· ·A·················· ····················· ···················· ···················· ····················
Q 2780 hCoV-19/USA/CA-ALSR-3053/2020|EPI_ISL_635817|2020- ···················· ·A·················· ····················· ···················· ···················· ····················
Q 2781 hCoV-19/USA/CA-ALSR-3054/2020|EPI_ISL_635818|2020- ···················· ···················· ····················· ···················· ···················· ····················
Q 2782 hCoV-19/USA/CA-ALSR-3057/2020|EPI_ISL_635819|2020- ···················· ·A·················· ····················· ···················· ···················· ····················
Q 2783 hCoV-19/USA/CA-ALSR-3058/2020|EPI_ISL_635820|2020- ···················· ···················· ····················· ···················· ···················· ····················
Q 2784 hCoV-19/USA/CA-ALSR-3059/2020|EPI_ISL_635821|2020- ···················· ···················· ····················· ···················· ···················· ····················
Q 2785 hCoV-19/USA/CA-ALSR-3060/2020|EPI_ISL_635822|2020- ···················· ·A·················· ····················· ···················· ···················· ····················
Q 2786 hCoV-19/USA/CA-ALSR-3062/2020|EPI_ISL_635823|2020- ···················· ·A·················· ····················· ···················· ···················· ····················
Q 2787 hCoV-19/USA/CA-ALSR-3065/2020|EPI_ISL_635824|2020- ···················· ·A·················· ····················· ···················· ···················· ····················
Q 2788 hCoV-19/USA/CA-ALSR-3066/2020|EPI_ISL_635825|2020- ···················· ·A·················· ····················· ···················· ···················· ····················
Q 2789 hCoV-19/USA/CA-ALSR-3067/2020|EPI_ISL_635826|2020- ···················· ·A·················· ····················· ···················· ···················· ····················
Q 2790 hCoV-19/USA/CA-ALSR-3068/2020|EPI_ISL_635827|2020- ···················· ···················· ····················· ···················· ···················· ····················
Q 2791 hCoV-19/USA/CA-ALSR-3071/2020|EPI_ISL_635828|2020- ···················· ·A·················· ····················· ···················· ···················· ····················
Q 2792 hCoV-19/USA/CA-ALSR-3074/2020|EPI_ISL_635829|2020- ···················· ·A·················· ····················· ···················· ···················· ····················
Q 2793 hCoV-19/USA/CA-ALSR-3076/2020|EPI_ISL_635830|2020- ···················· ·A·················· ····················· ···················· ···················· ····················
Q 2794 hCoV-19/USA/CA-ALSR-3079/2020|EPI_ISL_635832|2020- ···················· ·A·················· ····················· ···················· ···················· ····················
Q 2795 hCoV-19/USA/CA-ALSR-3081/2020|EPI_ISL_635833|2020- ···················· ·A·················· ····················· ···················· ···················· ····················
Q 2796 hCoV-19/USA/CA-ALSR-3082/2020|EPI_ISL_635834|2020- ···················· ···················· ····················· ···················· ···················· ····················
Q 2797 hCoV-19/USA/CA-ALSR-3083/2020|EPI_ISL_635835|2020- ···················· ·A·················· ····················· ···················· ···················· ····················
Q 2798 hCoV-19/USA/CA-ALSR-3084/2020|EPI_ISL_635836|2020- ···················· ·A·················· ····················· ···················· ···················· ····················
Q 2799 hCoV-19/USA/CA-ALSR-3085/2020|EPI_ISL_635837|2020- ···················· ·A·················· ····················· ···················· ···················· ····················
Q 2800 hCoV-19/USA/CA-ALSR-3086/2020|EPI_ISL_635838|2020- ···················· ·A·················· ····················· ···················· ···················· ····················
Q 2801 hCoV-19/USA/CA-ALSR-3088/2020|EPI_ISL_635840|2020- ···················· ·A·················· ····················· ···················· ···················· ····················
Q 2802 hCoV-19/USA/CA-ALSR-3089/2020|EPI_ISL_635841|2020- ···················· ·A·················· ····················· ···················· ···················· ····················
Q 2803 hCoV-19/USA/CA-ALSR-3090/2020|EPI_ISL_635842|2020- ···················· ·A·················· ····················· ···················· ···················· ····················
Q 2804 hCoV-19/USA/CA-ALSR-3091/2020|EPI_ISL_635843|2020- ···················· ·A·················· ····················· ···················· ···················· ····················
Q 2805 hCoV-19/USA/CA-ALSR-3092/2020|EPI_ISL_635844|2020- ···················· ·A·················· ····················· ···················· ···················· ····················
Q 2806 hCoV-19/USA/CA-ALSR-3093/2020|EPI_ISL_635845|2020- ···················· ···················· ····················· ···················· ···················· ····················
Q 2807 hCoV-19/USA/CA-ALSR-3162/2020|EPI_ISL_635890|2020- ···················· ···················· ····················· ···················· ···················· ····················

| | | | | | | | |
|---|---|---|---|---|---|---|---|
| Q | 2808 hCoV-19/USA/CA-ALSR-3163/2020\|EPI_ISL_635891\|2020- | ···················· | ···················· | ···················· | ···················· | ···················· | ···················· |
| Q | 2809 hCoV-19/USA/CA-ALSR-3164/2020\|EPI_ISL_635892\|2020- | ···················· | ·A·················· | ···················· | ···················· | ···················· | ···················· |
| Q | 2810 hCoV-19/USA/CA-ALSR-3165/2020\|EPI_ISL_635893\|2020- | ···················· | ·A·················· | ···················· | ···················· | ···················· | ···················· |
| Q | 2811 hCoV-19/USA/CA-ALSR-3166/2020\|EPI_ISL_635894\|2020- | ···················· | ·A·················· | ···················· | ···················· | ···················· | ···················· |
| Q | 2812 hCoV-19/USA/CA-ALSR-3168/2020\|EPI_ISL_635895\|2020- | ···················· | ·A·················· | ···················· | ···················· | ···················· | ···················· |
| Q | 2813 hCoV-19/USA/CA-ALSR-3169/2020\|EPI_ISL_635896\|2020- | ···················· | ···················· | ···················· | ···················· | ···················· | ···················· |
| Q | 2814 hCoV-19/USA/CA-ALSR-3170/2020\|EPI_ISL_635897\|2020- | ···················· | ·A·················· | ···················· | ···················· | ···················· | ···················· |
| Q | 2815 hCoV-19/USA/CA-ALSR-3172/2020\|EPI_ISL_635898\|2020- | ···················· | ···················· | ···················· | ···················· | ···················· | ···················· |
| Q | 2816 hCoV-19/USA/CA-ALSR-3173/2020\|EPI_ISL_635899\|2020- | ···················· | ···················· | ···················· | ···················· | ···················· | ···················· |
| Q | 2818 hCoV-19/Wales/PHWC-484B2D/2020\|EPI_ISL_627301\|2020 | ···················· | ···················· | ···················· | ···················· | ···················· | ···················· |
| Q | 2819 hCoV-19/England/NOTT-1186D5/2020\|EPI_ISL_627309\|20 | ···················· | ···················· | ···················· | ···················· | ···················· | ···················· |
| Q | 2820 hCoV-19/England/QEUH-A4D46B/2020\|EPI_ISL_628600\|20 | ···················· | ···················· | ···················· | ···················· | ···················· | ···················· |
| Q | 2821 hCoV-19/England/QEUH-A4D489/2020\|EPI_ISL_628601\|20 | ···················· | ·A·················· | ···················· | ···················· | ···················· | ···················· |
| Q | 2822 hCoV-19/England/QEUH-A4D4E3/2020\|EPI_ISL_628602\|20 | ···················· | ···················· | ···················· | ···················· | ···················· | ···················· |
| Q | 2823 hCoV-19/England/QEUH-A4D54A/2020\|EPI_ISL_628603\|20 | ···················· | ···················· | ···················· | ···················· | ···················· | ···················· |
| Q | 2824 hCoV-19/England/QEUH-A4D577/2020\|EPI_ISL_628604\|20 | ···················· | ···················· | ···················· | ···················· | ······T············· | ···················· |
| Q | 2825 hCoV-19/England/QEUH-A4D5B3/2020\|EPI_ISL_628605\|20 | ···················· | ···················· | ···················· | ···················· | ···················· | ···················· |
| Q | 2826 hCoV-19/England/QEUH-A4D5FF/2020\|EPI_ISL_628606\|20 | ···················· | ···················· | ···················· | ···················· | ······T············· | ···················· |
| Q | 2827 hCoV-19/England/QEUH-A4D60B/2020\|EPI_ISL_628607\|20 | ···················· | ···················· | ···················· | ···················· | ···················· | ···················· |
| Q | 2828 hCoV-19/England/QEUH-A4D647/2020\|EPI_ISL_628608\|20 | ···················· | C··················· | ···················· | ···················· | ···················· | ···················· |
| Q | 2829 hCoV-19/England/QEUH-A4D726/2020\|EPI_ISL_628609\|20 | ···················· | ···················· | ···················· | ···················· | ···················· | ···················· |
| Q | 2830 hCoV-19/England/QEUH-A4D735/2020\|EPI_ISL_628610\|20 | ···················· | ···················· | ···················· | ···················· | ···················· | ···················· |
| Q | 2831 hCoV-19/England/QEUH-A4D771/2020\|EPI_ISL_628611\|20 | ···················· | ···················· | ···················· | ···················· | ···················· | ···················· |
| Q | 2832 hCoV-19/England/QEUH-A4D823/2020\|EPI_ISL_628612\|20 | ···················· | ···················· | ···················· | ···················· | ···················· | ···················· |
| Q | 2833 hCoV-19/England/QEUH-A4D832/2020\|EPI_ISL_628613\|20 | ···················· | ···················· | ···················· | ···················· | ···················· | ···················· |
| Q | 2834 hCoV-19/England/QEUH-A4D8D8/2020\|EPI_ISL_628614\|20 | ···················· | ···················· | ···················· | ···················· | ···················· | ···················· |
| Q | 2835 hCoV-19/England/QEUH-A4D920/2020\|EPI_ISL_628615\|20 | ···················· | ···················· | ···················· | ···················· | ···················· | ···················· |
| Q | 2836 hCoV-19/England/QEUH-A4DB2A/2020\|EPI_ISL_628616\|20 | ···················· | ···················· | ···················· | ···················· | ···················· | ···················· |
| Q | 2837 hCoV-19/England/QEUH-A4DB66/2020\|EPI_ISL_628617\|20 | ···················· | ···················· | ···················· | ···················· | ···················· | ···················· |
| Q | 2838 hCoV-19/England/QEUH-A4DBB1/2020\|EPI_ISL_628618\|20 | ···················· | ······C············· | ···················· | ···················· | ···················· | ···················· |
| Q | 2839 hCoV-19/England/QEUH-A614BC/2020\|EPI_ISL_628619\|20 | ···················· | ···················· | ···················· | ···················· | ···················· | ···················· |
| Q | 2840 hCoV-19/England/QEUH-A615C8/2020\|EPI_ISL_628620\|20 | ···················· | ···················· | ···················· | ···················· | ···················· | ···················· |
| Q | 2841 hCoV-19/England/QEUH-A615E6/2020\|EPI_ISL_628621\|20 | ···················· | ···················· | ···················· | ···················· | ···················· | ···················· |
| Q | 2842 hCoV-19/England/QEUH-A61601/2020\|EPI_ISL_628622\|20 | ···················· | ···················· | ···················· | ···················· | ···················· | ···················· |
| Q | 2843 hCoV-19/England/QEUH-A61610/2020\|EPI_ISL_628623\|20 | ···················· | ·A·················· | ···················· | ···················· | ···················· | ···A················ |
| Q | 2844 hCoV-19/England/QEUH-A6162F/2020\|EPI_ISL_628624\|20 | ···················· | ···················· | ···················· | ···················· | ···················· | ···················· |
| Q | 2845 hCoV-19/England/QEUH-A6164D/2020\|EPI_ISL_628625\|20 | ···················· | ···················· | ···················· | ···················· | ···················· | ···················· |
| Q | 2846 hCoV-19/Wales/QEUH-A6165C/2020\|EPI_ISL_628626\|2020 | ···················· | ···················· | ···················· | ···················· | ···················· | ···················· |
| Q | 2847 hCoV-19/England/QEUH-A6166B/2020\|EPI_ISL_628627\|20 | ···················· | ·A·················· | ···················· | ···················· | ···················· | ···A················ |
| Q | 2848 hCoV-19/England/QEUH-A616F2/2020\|EPI_ISL_628628\|20 | ···················· | ···················· | ···················· | ···················· | ···················· | ···················· |
| Q | 2849 hCoV-19/Scotland/QEUH-A6170E/2020\|EPI_ISL_628629\|2 | ···················· | ···················· | ···················· | ···················· | ···················· | ···················· |
| Q | 2850 hCoV-19/England/QEUH-A6174A/2020\|EPI_ISL_628630\|20 | ···················· | ···················· | ···················· | ···················· | ······T············· | ···················· |
| Q | 2851 hCoV-19/England/QEUH-A61768/2020\|EPI_ISL_628631\|20 | ···················· | ···················· | ···················· | ···················· | ···················· | ···················· |
| Q | 2852 hCoV-19/England/QEUH-A61777/2020\|EPI_ISL_628632\|20 | ···················· | ···················· | ···················· | ···················· | ···················· | ···················· |
| Q | 2853 hCoV-19/England/QEUH-A61795/2020\|EPI_ISL_628633\|20 | ···················· | ···················· | ···················· | ···················· | ···················· | ···················· |
| Q | 2854 hCoV-19/England/QEUH-A6181A/2020\|EPI_ISL_628634\|20 | ···················· | ···················· | ···················· | ···················· | ···················· | ···················· |
| Q | 2855 hCoV-19/England/QEUH-A61838/2020\|EPI_ISL_628635\|20 | ···················· | ···················· | ···················· | ···················· | ···················· | ···················· |
| Q | 2856 hCoV-19/England/QEUH-A61874/2020\|EPI_ISL_628636\|20 | ···················· | ···················· | ···················· | ···················· | ···················· | ···················· |
| Q | 2857 hCoV-19/England/QEUH-A61892/2020\|EPI_ISL_628637\|20 | ···················· | ···················· | ···················· | ···················· | ···················· | ···················· |
| Q | 2858 hCoV-19/England/QEUH-A618A1/2020\|EPI_ISL_628638\|20 | ···················· | ···················· | ···················· | ···················· | ···················· | ···················· |
| Q | 2859 hCoV-19/England/QEUH-A618B0/2020\|EPI_ISL_628639\|20 | ···················· | ·A·················· | ···················· | ···················· | ···················· | ···A················ |
| Q | 2860 hCoV-19/England/QEUH-AD0F20/2020\|EPI_ISL_629900\|20 | ···················· | ···················· | ···················· | ···················· | ···················· | ···················· |
| Q | 2861 hCoV-19/England/QEUH-AD2920/2020\|EPI_ISL_629901\|20 | ···················· | ···················· | ···················· | ···················· | ···················· | ···················· |
| Q | 2862 hCoV-19/England/QEUH-AD0940/2020\|EPI_ISL_629903\|20 | ···················· | ···················· | ···················· | ···················· | ···················· | ···················· |
| Q | 2863 hCoV-19/England/QEUH-AD1DAD/2020\|EPI_ISL_629904\|20 | ···················· | ···················· | ···················· | ···················· | ···················· | ···················· |
| Q | 2864 hCoV-19/England/QEUH-AD1CA0/2020\|EPI_ISL_629905\|20 | ···················· | ···················· | ···················· | ···················· | ···················· | ···················· |
| Q | 2865 hCoV-19/England/QEUH-AD62F4/2020\|EPI_ISL_629928\|20 | ···················· | ···················· | ···················· | ···················· | ···················· | ···················· |
| Q | 2866 hCoV-19/England/QEUH-AD2674/2020\|EPI_ISL_629929\|20 | ···················· | ···················· | ···················· | ···················· | ···················· | ···················· |
| Q | 2867 hCoV-19/England/QEUH-AD2735/2020\|EPI_ISL_629930\|20 | ···················· | ···················· | ···················· | ···················· | ···················· | ···················· |
| Q | 2868 hCoV-19/England/QEUH-AD3ED5/2020\|EPI_ISL_629931\|20 | ···················· | ···················· | ···················· | ···················· | ······T············· | ···················· |
| Q | 2869 hCoV-19/England/QEUH-AD0F3F/2020\|EPI_ISL_629933\|20 | ···················· | ···················· | ···················· | ···················· | ···················· | ···················· |
| Q | 2870 hCoV-19/England/QEUH-AD0EBA/2020\|EPI_ISL_629934\|20 | ···················· | ···················· | ···················· | ···················· | ···················· | ···················· |
| Q | 2871 hCoV-19/England/QEUH-AD1D9E/2020\|EPI_ISL_629935\|20 | ···················· | ···················· | ···················· | ···················· | ···················· | ···················· |
| Q | 2872 hCoV-19/England/QEUH-AD0685/2020\|EPI_ISL_629936\|20 | ···················· | ···················· | ···················· | ···················· | ···················· | ···················· |
| Q | 2873 hCoV-19/England/QEUH-AD1E13/2020\|EPI_ISL_629937\|20 | ···················· | ···················· | ···················· | ···················· | ···················· | ···················· |
| Q | 2874 hCoV-19/England/QEUH-AD2243/2020\|EPI_ISL_629938\|20 | ···················· | ···················· | ···················· | ···················· | ···················· | ···················· |
| Q | 2875 hCoV-19/England/QEUH-AD631F/2020\|EPI_ISL_629939\|20 | ···················· | ···················· | ···················· | ···················· | ···················· | ···················· |
| Q | 2876 hCoV-19/England/QEUH-AD2FA6/2020\|EPI_ISL_629941\|20 | ···················· | ···················· | ···················· | ···················· | ···················· | ···················· |
| Q | 2877 hCoV-19/England/QEUH-AD342E/2020\|EPI_ISL_629943\|20 | ···················· | ···················· | ···················· | ···················· | ···················· | ···················· |
| Q | 2878 hCoV-19/England/QEUH-AD188E/2020\|EPI_ISL_629944\|20 | ···················· | ···················· | ···················· | ···················· | ···················· | ···················· |
| Q | 2879 hCoV-19/England/QEUH-AD26CF/2020\|EPI_ISL_629945\|20 | ···················· | ···················· | ···················· | ···················· | ···················· | ···················· |
| Q | 2880 hCoV-19/England/QEUH-AD2FF1/2020\|EPI_ISL_629946\|20 | ···················· | ···················· | ···················· | ···················· | ···················· | ···················· |
| Q | 2881 hCoV-19/England/QEUH-AD0F5D/2020\|EPI_ISL_629947\|20 | ···················· | ···················· | ···················· | ···················· | ···················· | ···················· |
| Q | 2882 hCoV-19/England/QEUH-AD39C5/2020\|EPI_ISL_629948\|20 | ···················· | ···················· | ···················· | ···················· | ···················· | ···················· |
| Q | 2883 hCoV-19/England/QEUH-AD1D70/2020\|EPI_ISL_629949\|20 | ···················· | ···················· | ···················· | ···················· | ···················· | ···················· |
| Q | 2884 hCoV-19/England/QEUH-AD2568/2020\|EPI_ISL_629950\|20 | ···················· | ···················· | ···················· | ···················· | ···················· | ···················· |
| Q | 2885 hCoV-19/England/QEUH-AD0E9C/2020\|EPI_ISL_629951\|20 | ···················· | ···················· | ···················· | ···················· | ···················· | ···················· |
| Q | 2886 hCoV-19/England/QEUH-AD03BB/2020\|EPI_ISL_629952\|20 | ···················· | ···················· | ···················· | ···················· | ···················· | ···················· |
| Q | 2887 hCoV-19/England/QEUH-AD235F/2020\|EPI_ISL_629954\|20 | ···················· | ···················· | ···················· | ···················· | ···················· | ···················· |
| Q | 2888 hCoV-19/England/QEUH-AD3A4A/2020\|EPI_ISL_629955\|20 | ···················· | ···················· | ···················· | ···················· | ···················· | ···················· |

```
Q 2889 hCoV-19/England/QEUH-AD0ED8/2020|EPI_ISL_629956|20 ···················· ···················· ····················· ···················· ···················· ····················
Q 2890 hCoV-19/England/QEUH-AD2EA9/2020|EPI_ISL_629957|20 ···················· ···················· ····················· ···················· ···················· ····················
Q 2891 hCoV-19/England/QEUH-AD2D8E/2020|EPI_ISL_629958|20 ···················· ···················· ····················· ···················· ···················· ····················
Q 2892 hCoV-19/England/QEUH-AD1C73/2020|EPI_ISL_629959|20 ···················· ···················· ····················· ···················· ···················· ····················
Q 2893 hCoV-19/England/QEUH-AD0EAB/2020|EPI_ISL_629960|20 ···················· ···················· ····················· ···················· ···················· ····················
Q 2894 hCoV-19/England/QEUH-AD095F/2020|EPI_ISL_629961|20 ···················· ···················· ····················· ···················· ···················· ····················
Q 2895 hCoV-19/England/QEUH-AD3B1A/2020|EPI_ISL_629962|20 ···················· ···················· ····················· ···················· ···················· ····················
Q 2896 hCoV-19/England/QEUH-AD2DBB/2020|EPI_ISL_629963|20 ···················· ···················· ····················· ···················· ···················· ····················
Q 2897 hCoV-19/England/QEUH-AD1DCB/2020|EPI_ISL_629964|20 ···················· ···················· ····················· ···················· ···················· ····················
Q 2898 hCoV-19/England/QEUH-AD2726/2020|EPI_ISL_629965|20 ···················· ···················· ····················· ···················· ···················· ····················
Q 2899 hCoV-19/England/QEUH-AD2D24/2020|EPI_ISL_629966|20 ···················· ···················· ····················· ···················· ···················· ··A·················
Q 2900 hCoV-19/England/QEUH-AD2656/2020|EPI_ISL_629967|20 ···················· ···················· ····················· ···················· ···················· ····················
Q 2901 hCoV-19/England/QEUH-AD0EE7/2020|EPI_ISL_629968|20 ···················· ···················· ····················· ···················· ···················· ····················
Q 2902 hCoV-19/England/QEUH-AD2270/2020|EPI_ISL_629969|20 ···················· ···················· ····················· ···················· ···················· ····················
Q 2903 hCoV-19/England/ALDP-AA405F/2020|EPI_ISL_630910|20 ···················· ···················· ····················· ···················· ···················· ····················
Q 2904 hCoV-19/England/ALDP-AA5030/2020|EPI_ISL_630912|20 ···················· ···················· ····················· ···················· ···················· ····················
Q 2905 hCoV-19/England/ALDP-AA4FBC/2020|EPI_ISL_630913|20 ···················· ···················· ····················· ···················· ···················· ····················
Q 2906 hCoV-19/England/QEUH-AAF27C/2020|EPI_ISL_630915|20 ···················· ···················· ····················· ···················· ···················· ····················
Q 2907 hCoV-19/England/ALDP-AA1B8E/2020|EPI_ISL_630916|20 ···················· ···················· ····················· ···················· ···················· ····················
Q 2908 hCoV-19/England/ALDP-AA4CE2/2020|EPI_ISL_630917|20 ···················· ···················· ····················· ···················· ···················· ····················
Q 2909 hCoV-19/England/ALDP-AA4C6A/2020|EPI_ISL_630918|20 ···················· ···················· ····················· ···················· ···················· ····················
Q 2910 hCoV-19/England/ALDP-AA09C1/2020|EPI_ISL_630920|20 ···················· ···················· ····················· ···················· ···················· ····················
Q 2911 hCoV-19/England/ALDP-AA3964/2020|EPI_ISL_630921|20 ···················· ···················· ····················· ···················· ···················· ··A·················
Q 2912 hCoV-19/England/ALDP-AA1C21/2020|EPI_ISL_630922|20 ···················· ···················· ····················· ···················· ···················· ····················
Q 2913 hCoV-19/England/QEUH-AAE7AB/2020|EPI_ISL_630923|20 ···················· ···················· ····················· ···················· ···················· ····················
Q 2914 hCoV-19/England/ALDP-AA1B33/2020|EPI_ISL_630924|20 ···················· ···················· ····················· ···················· ···················· ····················
Q 2915 hCoV-19/England/ALDP-AA38D0/2020|EPI_ISL_630925|20 ···················· ···················· ····················· ···················· ···················· ····················
Q 2916 hCoV-19/England/QEUH-AAE750/2020|EPI_ISL_630926|20 ···················· ·A·················· ····················· ···················· ···················· ····················
Q 2917 hCoV-19/England/ALDP-AA5003/2020|EPI_ISL_630927|20 ···················· ···················· ····················· ···················· ···················· ····················
Q 2918 hCoV-19/England/ALDP-AA28D1/2020|EPI_ISL_630928|20 ···················· ···················· ····················· ···················· ···················· ····················
Q 2919 hCoV-19/England/ALDP-AA4B12/2020|EPI_ISL_630929|20 ···················· ···················· ····················· ···················· ···················· ····················
Q 2920 hCoV-19/England/ALDP-AA265F/2020|EPI_ISL_630930|20 ···················· ···················· ····················· ···················· ···················· ····················
Q 2921 hCoV-19/England/QEUH-AAE653/2020|EPI_ISL_630931|20 ···················· ·A·················· ····················· ···················· ···················· ····················
Q 2922 hCoV-19/England/ALDP-AA4FCB/2020|EPI_ISL_630932|20 ···················· ···················· ····················· ···················· ···················· ····················
Q 2923 hCoV-19/England/QEUH-AAE6EA/2020|EPI_ISL_630933|20 ···················· ···················· ····················· ···················· ···················· ····················
Q 2924 hCoV-19/England/ALDP-AA4DEF/2020|EPI_ISL_630934|20 ···················· ···················· ····················· ···················· ···················· ····················
Q 2925 hCoV-19/England/ALDP-AA1BD9/2020|EPI_ISL_630935|20 ···················· ···················· ····················· ···················· ···················· ····················
Q 2926 hCoV-19/England/ALDP-AA274D/2020|EPI_ISL_630938|20 ···················· ···················· ····················· ···················· ···················· ····················
Q 2927 hCoV-19/England/ALDP-AA4D85/2020|EPI_ISL_630939|20 ···················· ···················· ····················· ···················· ···················· ····················
Q 2928 hCoV-19/England/ALDP-AA4A9D/2020|EPI_ISL_630940|20 ···················· ·A·················· ····················· ···················· ···················· ····················
Q 2929 hCoV-19/England/QEUH-AAE6DB/2020|EPI_ISL_630941|20 ···················· ·A·················· ····················· ···················· ···················· ····················
Q 2930 hCoV-19/England/ALDP-AA4F16/2020|EPI_ISL_630942|20 ···················· ···················· ····················· ···················· ···················· ····················
Q 2931 hCoV-19/England/ALDP-AA506D/2020|EPI_ISL_630943|20 ···················· ···················· ····················· ···················· ···················· ····················
Q 2932 hCoV-19/England/ALDP-AA4F61/2020|EPI_ISL_630944|20 ···················· ···················· ····················· ···················· ···················· ····················
Q 2933 hCoV-19/Wales/ALDP-AA4D58/2020|EPI_ISL_630945|2020 ···················· ···················· ····················· ···················· ···················· ····················
Q 2934 hCoV-19/England/ALDP-AA3FBD/2020|EPI_ISL_630946|20 ···················· ···················· ····················· ···················· ···················· ····················
Q 2935 hCoV-19/England/ALDP-AA2A35/2020|EPI_ISL_630947|20 ···················· ···················· ····················· ···················· ···················· ····················
Q 2936 hCoV-19/England/QEUH-AAE7D8/2020|EPI_ISL_630948|20 ···················· ···················· ····················· ···················· ···················· ····················
Q 2937 hCoV-19/England/QEUH-AADFFF/2020|EPI_ISL_630949|20 ···················· ···················· ····················· ···················· ···················· ····················
Q 2938 hCoV-19/England/ALDP-AA4DFE/2020|EPI_ISL_630950|20 ···················· ···················· ····················· ···················· ···················· ····················
Q 2939 hCoV-19/England/CAMC-AEAAB9/2020|EPI_ISL_633150|20 ···················· ···················· ····················· ···················· ···················· ····················
Q 2940 hCoV-19/England/QEUH-AE509E/2020|EPI_ISL_633151|20 ···················· ···················· ····················· ···················· ······T············· ····················
Q 2941 hCoV-19/England/QEUH-AE4B34/2020|EPI_ISL_633152|20 ···················· ···················· ····················· ···················· ···················· ····················
Q 2942 hCoV-19/England/CAMC-AEAC95/2020|EPI_ISL_633153|20 ···················· ···················· ····················· ···················· ···················· ····················
Q 2943 hCoV-19/England/QEUH-AE5465/2020|EPI_ISL_633160|20 ···················· ···················· ····················· ···················· ···················· ····················
Q 2944 hCoV-19/England/CAMC-AEAA40/2020|EPI_ISL_633161|20 ···················· ···················· ····················· ···················· ···················· ····················
Q 2945 hCoV-19/England/QEUH-AE4A64/2020|EPI_ISL_633162|20 ···················· ···················· ····················· ···················· ···················· ····················
Q 2946 hCoV-19/England/CAMC-AEAB01/2020|EPI_ISL_633163|20 ···················· ···················· ····················· ···················· ···················· ····················
Q 2947 hCoV-19/England/CAMC-AEA99E/2020|EPI_ISL_633164|20 ···················· ···················· ····················· ···················· ······T············· ····················
Q 2948 hCoV-19/England/QEUH-AE541A/2020|EPI_ISL_633166|20 ···················· ···················· ····················· ···················· ···················· ····················
Q 2949 hCoV-19/England/QEUH-AE4E0E/2020|EPI_ISL_633167|20 ···················· ···················· ····················· ···················· ···················· ····················
Q 2950 hCoV-19/England/QEUH-AE53C2/2020|EPI_ISL_633168|20 ···················· ···················· ····················· ···················· ···················· ····················
Q 2951 hCoV-19/England/QEUH-AE5104/2020|EPI_ISL_633169|20 ···················· ···················· ····················· ···················· ···················· ····················
Q 2952 hCoV-19/England/QEUH-AE4E2C/2020|EPI_ISL_633190|20 ···················· ···················· ····················· ···················· ···················· ····················
Q 2953 hCoV-19/England/QEUH-AE4D4D/2020|EPI_ISL_633191|20 ···················· ···················· ····················· ···················· ···················· ····················
Q 2954 hCoV-19/England/CAMC-AEABA7/2020|EPI_ISL_633192|20 ···················· ···················· ····················· ···················· ···················· ····················
Q 2955 hCoV-19/England/QEUH-AE4F56/2020|EPI_ISL_633193|20 ···················· ···················· ····················· ···················· ···················· ····················
Q 2956 hCoV-19/England/CAMC-AEB159/2020|EPI_ISL_633194|20 ···················· ···················· ····················· ···················· ···················· ····················
Q 2957 hCoV-19/England/QEUH-AE5377/2020|EPI_ISL_633196|20 ···················· ···················· ····················· ···················· ···················· ····················
Q 2958 hCoV-19/England/CAMC-AEA8DD/2020|EPI_ISL_633197|20 ···················· ···················· ····················· ···················· ······T············· ····················
Q 2959 hCoV-19/England/CAMC-AEA837/2020|EPI_ISL_633199|20 ···················· ···················· ····················· ···················· ···················· ····················
Q 2960 hCoV-19/England/CAMC-AEA891/2020|EPI_ISL_633200|20 ···················· ···················· ····················· ···················· ···················· ····················
Q 2961 hCoV-19/England/QEUH-AE4F74/2020|EPI_ISL_633202|20 ···················· ···················· ····················· ···················· ···················· ····················
Q 2962 hCoV-19/England/QEUH-AE532C/2020|EPI_ISL_633203|20 ···················· ···················· ····················· ···················· ···················· ····················
Q 2963 hCoV-19/England/QEUH-AE4CD7/2020|EPI_ISL_633204|20 ···················· ···················· ····················· ···················· ···················· ····················
Q 2964 hCoV-19/England/CAMC-AEA943/2020|EPI_ISL_633205|20 ···················· ···················· ····················· ···················· ······T············· ····················
Q 2965 hCoV-19/England/QEUH-AE4C7D/2020|EPI_ISL_633206|20 ···················· ···················· ····················· ···················· ···················· ····················
Q 2966 hCoV-19/England/QEUH-AE4A19/2020|EPI_ISL_633207|20 ···················· ···················· ····················· ···················· ···················· ····················
Q 2967 hCoV-19/England/CAMC-AEAB6B/2020|EPI_ISL_633208|20 ···················· ···················· ····················· ···················· ···················· ····················
Q 2968 hCoV-19/England/QEUH-AE4976/2020|EPI_ISL_633209|20 ···················· ···················· ····················· ···················· ···················· ····················
```

| | | | | | | | |
|---|---|---|---|---|---|---|---|
| Q | 2969 hCoV-19/England/QEUH-AE49A3/2020\|EPI_ISL_633210\|20 | ···················· | ···················· | ····················· | ···················· | ···················· | ···················· |
| Q | 2970 hCoV-19/England/CAMC-AEAFF6/2020\|EPI_ISL_633211\|20 | ···················· | ···················· | ····················· | ···················· | ···················· | ···················· |
| Q | 2971 hCoV-19/England/CAMC-AEAEBD/2020\|EPI_ISL_633212\|20 | ···················· | ···················· | ····················· | ···················· | ···················· | ···················· |
| Q | 2972 hCoV-19/England/QEUH-AE51B9/2020\|EPI_ISL_633213\|20 | ···················· | ···················· | ····················· | ···················· | ···················· | ···················· |
| Q | 2973 hCoV-19/England/CAMC-AEAE08/2020\|EPI_ISL_633214\|20 | ···················· | ···················· | ····················· | ···················· | ···················· | ···················· |
| Q | 2974 hCoV-19/England/CAMC-AEACFF/2020\|EPI_ISL_633215\|20 | ···················· | ···················· | ····················· | ···················· | ···················· | ···················· |
| Q | 2975 hCoV-19/England/CAMC-AEA8EC/2020\|EPI_ISL_633216\|20 | ···················· | ···················· | ····················· | ···················· | ·····T·············· | ···················· |
| Q | 2976 hCoV-19/England/CAMC-AEACA4/2020\|EPI_ISL_633217\|20 | ···················· | ···················· | ····················· | ···················· | ···················· | ···················· |
| Q | 2977 hCoV-19/England/CAMC-AEABF2/2020\|EPI_ISL_633219\|20 | ···················· | ···················· | ····················· | ···················· | ···················· | ···················· |
| Q | 2978 hCoV-19/England/CAMC-AEB02F/2020\|EPI_ISL_633220\|20 | ···················· | ···················· | ····················· | ···················· | ···················· | ···················· |
| Q | 2979 hCoV-19/England/QEUH-AE4ACE/2020\|EPI_ISL_633222\|20 | ···················· | ···················· | ····················· | ···················· | ···················· | ···················· |
| Q | 2980 hCoV-19/England/CAMC-AEB1A4/2020\|EPI_ISL_633223\|20 | ···················· | ···················· | ····················· | ···················· | ···················· | ···················· |
| Q | 2981 hCoV-19/England/CAMC-AEAB5C/2020\|EPI_ISL_633224\|20 | ···················· | ···················· | ····················· | ···················· | ···················· | ···················· |
| Q | 2982 hCoV-19/England/CAMC-AEA679/2020\|EPI_ISL_633225\|20 | ···················· | ···················· | ····················· | ···················· | ·····T·············· | ···················· |
| Q | 2983 hCoV-19/England/QEUH-AE4B70/2020\|EPI_ISL_633226\|20 | ···················· | ···················· | ····················· | ···················· | ···················· | ···················· |
| Q | 2984 hCoV-19/England/CAMC-AEB11D/2020\|EPI_ISL_633227\|20 | ···················· | ···················· | ····················· | ···················· | ···················· | ···················· |
| Q | 2985 hCoV-19/England/MILK-ACD23C/2020\|EPI_ISL_634482\|20 | ···················· | ···················· | ····················· | ···················· | ···················· | ···················· |
| Q | 2986 hCoV-19/England/MILK-AC78E5/2020\|EPI_ISL_634484\|20 | ···················· | ···················· | ····················· | ···················· | ···················· | ···················· |
| Q | 2987 hCoV-19/England/MILK-ACD041/2020\|EPI_ISL_634490\|20 | ···················· | ···················· | ····················· | ···················· | ···T················ | ···················· |
| Q | 2988 hCoV-19/England/MILK-AC788B/2020\|EPI_ISL_634491\|20 | ···················· | ·A·················· | ····················· | ···················· | ···················· | ··A················· |
| Q | 2989 hCoV-19/England/MILK-AC7BFB/2020\|EPI_ISL_634494\|20 | ···················· | ···················· | ····················· | ···················· | ···················· | ···················· |
| Q | 2990 hCoV-19/England/MILK-AC79B5/2020\|EPI_ISL_634495\|20 | ···················· | ···················· | ····················· | ···················· | ···················· | ···················· |
| Q | 2991 hCoV-19/England/MILK-ACD20F/2020\|EPI_ISL_634497\|20 | ···················· | ···················· | ····················· | ···················· | ···················· | ···················· |
| Q | 2992 hCoV-19/England/MILK-AD012A/2020\|EPI_ISL_634498\|20 | ···················· | ···················· | ····················· | ···················· | ···················· | ···················· |
| Q | 2993 hCoV-19/England/MILK-ACDA16/2020\|EPI_ISL_634515\|20 | ···················· | ···················· | ····················· | ···················· | ···················· | ···················· |
| Q | 2994 hCoV-19/England/MILK-ACC516/2020\|EPI_ISL_634516\|20 | ···················· | ···················· | ····················· | ···················· | ···················· | ···················· |
| Q | 2995 hCoV-19/England/MILK-AC8107/2020\|EPI_ISL_634518\|20 | ···················· | ···················· | ····················· | ···················· | ·····T·············· | ···················· |
| Q | 2996 hCoV-19/England/MILK-ACD418/2020\|EPI_ISL_634519\|20 | ···················· | ···················· | ····················· | ···················· | ···················· | ···················· |
| Q | 2997 hCoV-19/England/QEUH-AD438A/2020\|EPI_ISL_634520\|20 | ···················· | ···················· | ····················· | ···················· | ···················· | ···················· |
| Q | 2998 hCoV-19/England/QEUH-AD43E4/2020\|EPI_ISL_634521\|20 | ···················· | ···················· | ····················· | ···················· | ···················· | ···················· |
| Q | 2999 hCoV-19/England/QEUH-AD5486/2020\|EPI_ISL_634522\|20 | ···················· | ···················· | ····················· | ···················· | ···················· | ···················· |
| Q | 3000 hCoV-19/England/QEUH-AD4C34/2020\|EPI_ISL_634523\|20 | ···················· | ···················· | ····················· | ···················· | ···················· | ···················· |
| Q | 3001 hCoV-19/England/QEUH-AD5732/2020\|EPI_ISL_634524\|20 | ···················· | ·A·················· | ····················· | ···················· | ···················· | ··A················· |
| Q | 3002 hCoV-19/England/QEUH-AD46FA/2020\|EPI_ISL_634525\|20 | ···················· | ···················· | ····················· | ···················· | ···················· | ···················· |
| Q | 3003 hCoV-19/England/QEUH-AD1F2F/2020\|EPI_ISL_634526\|20 | ···················· | ···················· | ····················· | ···················· | ···················· | ···················· |
| Q | 3004 hCoV-19/England/QEUH-AD43F3/2020\|EPI_ISL_634527\|20 | ···················· | ···················· | ····················· | ···················· | ···················· | ···················· |
| Q | 3005 hCoV-19/England/QEUH-AD4029/2020\|EPI_ISL_634528\|20 | ···················· | ···················· | ····················· | ···················· | ···················· | ···················· |
| Q | 3006 hCoV-19/England/QEUH-AD5C42/2020\|EPI_ISL_634529\|20 | ···················· | ···················· | ····················· | ···················· | ···················· | ···················· |
| Q | 3007 hCoV-19/England/QEUH-AD445A/2020\|EPI_ISL_634530\|20 | ···················· | ···················· | ····················· | ···················· | ···················· | ···················· |
| Q | 3008 hCoV-19/England/QEUH-AD5CD9/2020\|EPI_ISL_634531\|20 | ···················· | ···················· | ····················· | ···················· | ·····T·············· | ···················· |
| Q | 3009 hCoV-19/England/QEUH-AD3F5A/2020\|EPI_ISL_634532\|20 | ···················· | ···················· | ····················· | ···················· | ···················· | ···················· |
| Q | 3010 hCoV-19/England/QEUH-AD19B8/2020\|EPI_ISL_634533\|20 | ···················· | ···················· | ····················· | ···················· | ···················· | ···················· |
| Q | 3011 hCoV-19/England/QEUH-AD100E/2020\|EPI_ISL_634535\|20 | ···················· | ···················· | ····················· | ···················· | ···················· | ···················· |
| Q | 3012 hCoV-19/England/QEUH-AD1F5C/2020\|EPI_ISL_634536\|20 | ···················· | ···················· | ····················· | ···················· | ···················· | ···················· |
| Q | 3013 hCoV-19/England/QEUH-AD401A/2020\|EPI_ISL_634538\|20 | ···················· | ···················· | ····················· | ···················· | ···················· | ···················· |
| Q | 3014 hCoV-19/England/QEUH-AD16FD/2020\|EPI_ISL_634539\|20 | ···················· | ···················· | ····················· | ···················· | ···················· | ···················· |
| Q | 3015 hCoV-19/England/QEUH-AD5E00/2020\|EPI_ISL_634541\|20 | ···················· | ···················· | ····················· | ···················· | ···················· | ···················· |
| Q | 3016 hCoV-19/England/QEUH-AD5E97/2020\|EPI_ISL_634542\|20 | ···················· | ···················· | ····················· | ···················· | ···················· | ···················· |
| Q | 3017 hCoV-19/England/QEUH-AD5B90/2020\|EPI_ISL_634543\|20 | ···················· | ···················· | ····················· | ···················· | ···················· | ···················· |
| Q | 3018 hCoV-19/England/QEUH-AD58C6/2020\|EPI_ISL_634544\|20 | ···················· | ···················· | ····················· | ···················· | ···················· | ···················· |
| Q | 3019 hCoV-19/England/QEUH-AD4CAD/2020\|EPI_ISL_634545\|20 | ···················· | ···················· | ····················· | ···················· | ···················· | ···················· |
| Q | 3020 hCoV-19/England/QEUH-AD1059/2020\|EPI_ISL_634546\|20 | ···················· | ···················· | ····················· | ···················· | ···················· | ···················· |
| Q | 3021 hCoV-19/England/QEUH-AD198B/2020\|EPI_ISL_634547\|20 | ···················· | ···················· | ····················· | ···················· | ···················· | ···················· |
| Q | 3022 hCoV-19/England/QEUH-AD4C70/2020\|EPI_ISL_634548\|20 | ···················· | ···················· | ····················· | ···················· | ···················· | ···················· |
| Q | 3023 hCoV-19/England/QEUH-AD5A75/2020\|EPI_ISL_634549\|20 | ···················· | ···················· | ····················· | ···················· | ···················· | ···················· |
| Q | 3024 hCoV-19/England/QEUH-AD5F0D/2020\|EPI_ISL_634550\|20 | ···················· | ···················· | ····················· | ···················· | ···················· | ···················· |
| Q | 3025 hCoV-19/England/QEUH-AD3576/2020\|EPI_ISL_634551\|20 | ···················· | ···················· | ····················· | ···················· | ···················· | ···················· |
| Q | 3026 hCoV-19/England/QEUH-AD42BA/2020\|EPI_ISL_634552\|20 | ···················· | ···················· | ····················· | ···················· | ···················· | ···················· |
| Q | 3027 hCoV-19/England/QEUH-AD194F/2020\|EPI_ISL_634553\|20 | ···················· | ···················· | ····················· | ···················· | ···················· | ···················· |
| Q | 3028 hCoV-19/England/QEUH-AD5EE2/2020\|EPI_ISL_634554\|20 | ···················· | ·A·················· | ····················· | ···················· | ···················· | ··A················· |
| Q | 3029 hCoV-19/England/QEUH-AD4180/2020\|EPI_ISL_634555\|20 | ···················· | ···················· | ····················· | ···················· | ···················· | ···················· |
| Q | 3030 hCoV-19/England/QEUH-AD4047/2020\|EPI_ISL_634556\|20 | ···················· | ···················· | ····················· | ···················· | ···················· | ···················· |
| Q | 3031 hCoV-19/England/QEUH-AD4681/2020\|EPI_ISL_634557\|20 | ···················· | ···················· | ····················· | ···················· | ···················· | ···················· |
| Q | 3032 hCoV-19/England/QEUH-AD45FD/2020\|EPI_ISL_634558\|20 | ···················· | ···················· | ····················· | ···················· | ···················· | ···················· |
| Q | 3033 hCoV-19/USA/CA-ALSR-3095/2020\|EPI_ISL_635846\|2020- | ···················· | ···················· | ····················· | ···················· | ···················· | ···················· |
| Q | 3034 hCoV-19/USA/CA-ALSR-3096/2020\|EPI_ISL_635847\|2020- | ···················· | ·A·················· | ····················· | ···················· | ···················· | ···················· |
| Q | 3035 hCoV-19/USA/CA-ALSR-3097/2020\|EPI_ISL_635848\|2020- | ···················· | ···················· | ····················· | ···················· | ···················· | ···················· |
| Q | 3036 hCoV-19/USA/CA-ALSR-3099/2020\|EPI_ISL_635849\|2020- | ···················· | ···················· | ····················· | ···················· | ···················· | ···················· |
| Q | 3037 hCoV-19/USA/CA-ALSR-3106/2020\|EPI_ISL_635852\|2020- | ···················· | ···················· | ····················· | ···················· | ···················· | ···················· |
| Q | 3038 hCoV-19/USA/CA-ALSR-3111/2020\|EPI_ISL_635853\|2020- | ···················· | ···················· | ····················· | ···················· | ···················· | ···················· |
| Q | 3039 hCoV-19/USA/CA-ALSR-3112/2020\|EPI_ISL_635854\|2020- | ···················· | ·A·················· | ····················· | ···················· | ···················· | ···················· |
| Q | 3040 hCoV-19/USA/CA-ALSR-3117/2020\|EPI_ISL_635855\|2020- | ···················· | ···················· | ····················· | ···················· | ···················· | ···················· |
| Q | 3041 hCoV-19/USA/CA-ALSR-3118/2020\|EPI_ISL_635856\|2020- | ···················· | ···················· | ····················· | ···················· | ···················· | ···················· |
| Q | 3042 hCoV-19/USA/CA-ALSR-3119/2020\|EPI_ISL_635857\|2020- | ···················· | ·A·················· | ····················· | ···················· | ···················· | ···················· |
| Q | 3043 hCoV-19/USA/CA-ALSR-3120/2020\|EPI_ISL_635858\|2020- | ···················· | ·A·················· | ····················· | ···················· | ···················· | ···················· |
| Q | 3044 hCoV-19/USA/CA-ALSR-3126/2020\|EPI_ISL_635859\|2020- | ········T··········· | ·A·················· | ····················· | ···················· | ···················· | ···················· |
| Q | 3045 hCoV-19/USA/CA-ALSR-3127/2020\|EPI_ISL_635860\|2020- | ···················· | ·A····A············· | ····················· | ···················· | ···················· | ···················· |
| Q | 3046 hCoV-19/USA/CA-ALSR-3128/2020\|EPI_ISL_635861\|2020- | ···················· | ·A·················· | ····················· | ···················· | ···················· | ···················· |
| Q | 3047 hCoV-19/USA/CA-ALSR-3129/2020\|EPI_ISL_635862\|2020- | ···················· | ·A·················· | ····················· | ···················· | ···················· | ···················· |
| Q | 3048 hCoV-19/USA/CA-ALSR-3131/2020\|EPI_ISL_635863\|2020- | ···················· | ···················· | ····················· | ···················· | ···················· | ···················· |

Q 3049 hCoV-19/USA/CA-ALSR-3132/2020|EPI_ISL_635864|2020- ···················· ·A·················· ····················· ···················· ···················· ····················
Q 3050 hCoV-19/USA/CA-ALSR-3133/2020|EPI_ISL_635865|2020- ···················· ·A·················· ····················· ···················· ···················· ····················
Q 3051 hCoV-19/USA/CA-ALSR-3134/2020|EPI_ISL_635866|2020- ···················· ·A·················· ····················· ···················· ···················· ····················
Q 3052 hCoV-19/USA/CA-ALSR-3135/2020|EPI_ISL_635867|2020- ···················· ···················· ····················· ···················· ···················· ····················
Q 3053 hCoV-19/USA/CA-ALSR-3137/2020|EPI_ISL_635868|2020- ···················· ·A·················· ····················· ···················· ···················· ····················
Q 3054 hCoV-19/USA/CA-ALSR-3139/2020|EPI_ISL_635869|2020- ···················· ·A·················· ····················· ···················· ···················· ····················
Q 3055 hCoV-19/USA/CA-ALSR-3140/2020|EPI_ISL_635870|2020- ···················· ·A·················· ····················· ···················· ···················· ····················
Q 3056 hCoV-19/USA/CA-ALSR-3141/2020|EPI_ISL_635871|2020- ···················· ·A·················· ····················· ···················· ···················· ····················
Q 3057 hCoV-19/USA/CA-ALSR-3143/2020|EPI_ISL_635873|2020- ···················· ·A·················· ····················· ···················· ···················· ····················
Q 3058 hCoV-19/USA/CA-ALSR-3144/2020|EPI_ISL_635874|2020- ···················· ···················· ····················· ···················· ···················· ····················
Q 3059 hCoV-19/USA/CA-ALSR-3145/2020|EPI_ISL_635875|2020- ···················· ·A·················· ····················· ···················· ···················· ····················
Q 3060 hCoV-19/USA/CA-ALSR-3146/2020|EPI_ISL_635876|2020- ···················· ·A·················· ····················· ···················· ···················· ····················
Q 3061 hCoV-19/USA/CA-ALSR-3147/2020|EPI_ISL_635877|2020- ···················· ·A·················· ····················· ···················· ···················· ····················
Q 3062 hCoV-19/USA/CA-ALSR-3148/2020|EPI_ISL_635878|2020- ···················· ·A·················· ····················· ···················· ···················· ····················
Q 3063 hCoV-19/USA/CA-ALSR-3149/2020|EPI_ISL_635879|2020- ···················· ·A·················· ····················· ···················· ···················· ····················
Q 3064 hCoV-19/USA/CA-ALSR-3151/2020|EPI_ISL_635880|2020- ···················· ·A·················· ····················· ···················· ···················· ····················
Q 3065 hCoV-19/USA/CA-ALSR-3153/2020|EPI_ISL_635881|2020- ···················· ·A·················· ····················· ···················· ···················· ····················
Q 3066 hCoV-19/USA/CA-ALSR-3154/2020|EPI_ISL_635882|2020- ···················· ···················· ····················· ···················· ···················· ····················
Q 3067 hCoV-19/USA/CA-ALSR-3155/2020|EPI_ISL_635883|2020- ···················· ·A·················· ····················· ···················· ···················· ····················
Q 3068 hCoV-19/USA/CA-ALSR-3157/2020|EPI_ISL_635885|2020- ···················· ·A·················· ····················· ···················· ···················· ····················
Q 3069 hCoV-19/USA/CA-ALSR-3158/2020|EPI_ISL_635886|2020- ···················· ·A·················· ····················· ···················· ···················· ····················
Q 3070 hCoV-19/USA/CA-ALSR-3159/2020|EPI_ISL_635887|2020- ···················· ·A·················· ····················· ···················· ···················· ····················
Q 3071 hCoV-19/USA/CA-ALSR-3160/2020|EPI_ISL_635888|2020- ···················· ·A·················· ····················· ···················· ···················· ····················
Q 3072 hCoV-19/USA/CA-ALSR-3161/2020|EPI_ISL_635889|2020- ···················· ···················· ····················· ···················· ···················· ····················
Q 3074 hCoV-19/England/CAMC-A6603B/2020|EPI_ISL_627314|20 ···················· ···················· ····················· ···················· ···················· ····················
Q 3075 hCoV-19/England/CAMC-A66059/2020|EPI_ISL_627315|20 ···················· ···················· ····················· ···················· ···················· ····················
Q 3076 hCoV-19/England/CAMC-A66226/2020|EPI_ISL_627316|20 ···················· ···················· ····················· ···················· ···················· ····················
Q 3078 hCoV-19/England/CAMC-A663AB/2020|EPI_ISL_627318|20 ···················· ···················· ····················· ···················· ···················· ····················
Q 3079 hCoV-19/England/CAMC-A6651E/2020|EPI_ISL_627319|20 ···················· ···················· ····················· ···················· ···················· ····················
Q 3080 hCoV-19/England/CAMC-A665A5/2020|EPI_ISL_627320|20 ···················· ···················· ····················· ···················· ···················· ····················
Q 3081 hCoV-19/England/CAMC-A665E1/2020|EPI_ISL_627321|20 ···················· ···················· ····················· ···················· ···················· ····················
Q 3083 hCoV-19/England/CAMC-A666B1/2020|EPI_ISL_627323|20 ···················· ···················· ····················· ···················· ······T············· ····················
Q 3084 hCoV-19/England/CAMC-A666FD/2020|EPI_ISL_627324|20 ···················· ···················· ····················· ···················· ···················· ····················
Q 3085 hCoV-19/England/CAMC-A66709/2020|EPI_ISL_627325|20 ···················· ···················· ····················· ···················· ···················· ····················
Q 3086 hCoV-19/England/CAMC-A66718/2020|EPI_ISL_627326|20 ···················· ···················· ····················· ···················· ···················· ····················
Q 3087 hCoV-19/England/CAMC-A66736/2020|EPI_ISL_627327|20 ···················· ···················· ····················· ···················· ···················· ····················
Q 3088 hCoV-19/England/CAMC-A66790/2020|EPI_ISL_627328|20 ···················· ···················· ····················· ···················· ···················· ····················
Q 3089 hCoV-19/Wales/CVR4736/2020|EPI_ISL_627329|2020-09- ···················· ···················· ····················· ···················· ···················· ····················
Q 3090 hCoV-19/Wales/CVR4737/2020|EPI_ISL_627330|2020-09- ···················· ···················· ····················· ···················· ···················· ····················
Q 3091 hCoV-19/Wales/CVR4738/2020|EPI_ISL_627331|2020-09- ···················· ···················· ····················· ···················· ···················· ····················
Q 3092 hCoV-19/Wales/CVR4741/2020|EPI_ISL_627333|2020-09- ···················· ···················· ····················· ···················· ···················· ····················
Q 3093 hCoV-19/Wales/CVR4743/2020|EPI_ISL_627334|2020-09- ···················· ···················· ····················· ···················· ···················· ····················
Q 3094 hCoV-19/Wales/CVR4744/2020|EPI_ISL_627335|2020-09- ···················· ···················· ····················· ···················· ···················· ····················
Q 3095 hCoV-19/Wales/CVR4745/2020|EPI_ISL_627336|2020-09- ···················· ···················· ····················· ···················· ···················· ····················
Q 3096 hCoV-19/Wales/CVR4746/2020|EPI_ISL_627337|2020-09- ···················· ···················· ····················· ···················· ···················· ····················
Q 3097 hCoV-19/Wales/CVR4747/2020|EPI_ISL_627338|2020-09- ···················· ···················· ····················· ···················· ···················· ····················
Q 3098 hCoV-19/Wales/CVR4749/2020|EPI_ISL_627339|2020-09- ···················· ···················· ····················· ···················· ···················· ····················
Q 3099 hCoV-19/Wales/CVR4759/2020|EPI_ISL_627340|2020-09- ···················· ···················· ····················· ···················· ···················· ····················
Q 3100 hCoV-19/Wales/CVR4760/2020|EPI_ISL_627341|2020-09- ···················· ···················· ····················· ···················· ···················· ····················
Q 3101 hCoV-19/Wales/CVR4762/2020|EPI_ISL_627342|2020-09- ···················· ···················· ····················· ···················· ···················· ····················
Q 3102 hCoV-19/Wales/CVR4763/2020|EPI_ISL_627343|2020-09- ···················· ···················· ····················· ···················· ···················· ····················
Q 3103 hCoV-19/Wales/CVR4771/2020|EPI_ISL_627344|2020-09- ···················· ···················· ····················· ···················· ···················· ····················
Q 3104 hCoV-19/Wales/CVR4772/2020|EPI_ISL_627345|2020-09- ···················· ···················· ····················· ···················· ···················· ····················
Q 3105 hCoV-19/Scotland/CVR4775/2020|EPI_ISL_627346|2020- ···················· ···················· ····················· ···················· ···················· ····················
Q 3106 hCoV-19/Scotland/CVR4776/2020|EPI_ISL_627347|2020- ···················· ···················· ····················· ···················· ···················· ····················
Q 3107 hCoV-19/Scotland/CVR4778/2020|EPI_ISL_627348|2020- ···················· ···················· ····················· ···················· ···················· ····················
Q 3108 hCoV-19/Scotland/CVR4780/2020|EPI_ISL_627349|2020- ···················· ···················· ····················· ···················· ···················· ····················
Q 3109 hCoV-19/Wales/CVR4782/2020|EPI_ISL_627350|2020-09- ···················· ···················· ····················· ···················· ···················· ····················
Q 3110 hCoV-19/Wales/CVR4788/2020|EPI_ISL_627351|2020-09- ···················· ···················· ····················· ···················· ···················· ····················
Q 3111 hCoV-19/Wales/CVR4790/2020|EPI_ISL_627352|2020-09- ···················· ···················· ····················· ···················· ···················· ····················
Q 3112 hCoV-19/Wales/CVR4794/2020|EPI_ISL_627353|2020-09- ···················· ···················· ····················· ···················· ···················· ····················
Q 3113 hCoV-19/Wales/CVR4799/2020|EPI_ISL_627354|2020-09- ···················· ···················· ····················· ···················· ···················· ····················
Q 3114 hCoV-19/Wales/CVR4804/2020|EPI_ISL_627355|2020-09- ···················· ···················· ····················· ···················· ···················· ····················
Q 3115 hCoV-19/Wales/CVR4806/2020|EPI_ISL_627356|2020-09- ···················· ···················· ····················· ···················· ···················· ····················
Q 3116 hCoV-19/Wales/CVR4809/2020|EPI_ISL_627357|2020-09- ···················· ···················· ····················· ···················· ···················· ····················
Q 3117 hCoV-19/Wales/CVR4815/2020|EPI_ISL_627358|2020-09- ···················· ···················· ····················· ···················· ···················· ····················
Q 3118 hCoV-19/Wales/CVR4824/2020|EPI_ISL_627359|2020-09- ···················· ···················· ····················· ···················· ···················· ····················
Q 3119 hCoV-19/Wales/CVR4828/2020|EPI_ISL_627360|2020-09- ···················· ···················· ····················· ···················· ···················· ····················
Q 3120 hCoV-19/Scotland/CVR4835/2020|EPI_ISL_627361|2020- ···················· ···················· ····················· ···················· ···················· ····················
Q 3121 hCoV-19/Scotland/CVR4836/2020|EPI_ISL_627362|2020- ···················· ···················· ····················· ···················· ···················· ····················
Q 3122 hCoV-19/Scotland/CVR4837/2020|EPI_ISL_627363|2020- ···················· ···················· ····················· ···················· ···················· ····················
Q 3123 hCoV-19/Scotland/CVR4838/2020|EPI_ISL_627364|2020- ···················· ···················· ····················· ···················· ···················· ····················
Q 3124 hCoV-19/Scotland/CVR4839/2020|EPI_ISL_627365|2020- ···················· ···················· ····················· ···················· ···················· ····················
Q 3125 hCoV-19/Scotland/CVR4840/2020|EPI_ISL_627366|2020- ···················· ···················· ····················· ···················· ···················· ····················
Q 3126 hCoV-19/Scotland/CVR4841/2020|EPI_ISL_627367|2020- ···················· ···················· ····················· ···················· ···················· ····················
Q 3127 hCoV-19/Scotland/CVR5091/2020|EPI_ISL_627368|2020- ···················· ···················· ····················· ···················· ···················· ····················
Q 3128 hCoV-19/Scotland/CVR5093/2020|EPI_ISL_627369|2020- ···················· ···················· ····················· ···················· ···················· ····················
Q 3129 hCoV-19/Scotland/CVR5094/2020|EPI_ISL_627370|2020- ···················· ···················· ····················· ···················· ···················· ····················
Q 3130 hCoV-19/Scotland/CVR5095/2020|EPI_ISL_627371|2020- ···················· ···················· ····················· ···················· ···················· ····················
Q 3131 hCoV-19/Scotland/CVR5096/2020|EPI_ISL_627372|2020- ···················· ···················· ····················· ···················· ···················· ····················

| | | | | | | | |
|---|---|---|---|---|---|---|---|
| Q | 3132 hCoV-19/Scotland/CVR5104/2020\|EPI_ISL_627373\|2020- | ···················· | ···················· | ···················· | ···················· | ···················· | ···················· |
| Q | 3133 hCoV-19/Scotland/CVR5105/2020\|EPI_ISL_627374\|2020- | ···················· | ···················· | ···················· | ···················· | ···················· | ···················· |
| Q | 3134 hCoV-19/Scotland/CVR5106/2020\|EPI_ISL_627375\|2020- | ···················· | ···················· | ···················· | ···················· | ···················· | ···················· |
| Q | 3135 hCoV-19/Scotland/CVR5109/2020\|EPI_ISL_627376\|2020- | ···················· | ···················· | ···················· | ···················· | ···················· | ···················· |
| Q | 3136 hCoV-19/Scotland/CVR5113/2020\|EPI_ISL_627377\|2020- | ···················· | ···················· | ···················· | ···················· | ···················· | ···················· |
| Q | 3137 hCoV-19/Scotland/CVR5129/2020\|EPI_ISL_627378\|2020- | ···················· | ···················· | ···················· | ···················· | ···················· | ···················· |
| Q | 3138 hCoV-19/Scotland/CVR5140/2020\|EPI_ISL_627379\|2020- | ···················· | ···················· | ···················· | ···················· | ···················· | ···················· |
| Q | 3139 hCoV-19/Scotland/CVR5178/2020\|EPI_ISL_627380\|2020- | ···················· | ···················· | ···················· | ···················· | ···················· | ···················· |
| Q | 3141 hCoV-19/Scotland/CVR5185/2020\|EPI_ISL_627382\|2020- | ···················· | ···················· | ···················· | ···················· | ···················· | ···················· |
| Q | 3142 hCoV-19/Scotland/CVR5186/2020\|EPI_ISL_627383\|2020- | ···················· | ···················· | ···················· | ···················· | ···················· | ···················· |
| Q | 3143 hCoV-19/Scotland/CVR5190/2020\|EPI_ISL_627384\|2020- | ···················· | ···················· | ···················· | ···················· | ···················· | ···················· |
| Q | 3144 hCoV-19/Scotland/CVR5196/2020\|EPI_ISL_627385\|2020- | ···················· | ···················· | ···················· | ···················· | ···················· | ···················· |
| Q | 3145 hCoV-19/Scotland/CVR5197/2020\|EPI_ISL_627386\|2020- | ···················· | ···················· | ···················· | ···················· | ···················· | ···················· |
| Q | 3146 hCoV-19/Scotland/CVR5200/2020\|EPI_ISL_627387\|2020- | ···················· | ···················· | ···················· | ···················· | ···················· | ···················· |
| Q | 3147 hCoV-19/Scotland/CVR5202/2020\|EPI_ISL_627388\|2020- | ···················· | ···················· | ···················· | ···················· | ···················· | ···················· |
| Q | 3149 hCoV-19/England/QEUH-A618CF/2020\|EPI_ISL_628640\|20 | ···················· | ···················· | ···················· | ···················· | ···················· | ···················· |
| Q | 3150 hCoV-19/Scotland/QEUH-A618DE/2020\|EPI_ISL_628641\|2 | ···················· | ···················· | ···················· | ···················· | ···················· | ···················· |
| Q | 3151 hCoV-19/England/QEUH-A618FC/2020\|EPI_ISL_628642\|20 | ···················· | ···················· | ···················· | ···················· | ···················· | ···················· |
| Q | 3152 hCoV-19/England/QEUH-A61908/2020\|EPI_ISL_628643\|20 | ···················· | ···················· | ···················· | ···················· | ···················· | ···················· |
| Q | 3153 hCoV-19/England/QEUH-A61953/2020\|EPI_ISL_628644\|20 | ···················· | ···················· | ···················· | ···················· | ···················· | ···················· |
| Q | 3154 hCoV-19/England/QEUH-A61962/2020\|EPI_ISL_628645\|20 | ···················· | ···················· | ···················· | ···················· | ···················· | ···················· |
| Q | 3155 hCoV-19/England/QEUH-A61A14/2020\|EPI_ISL_628646\|20 | ···················· | ···················· | ···················· | ···················· | ····T··············· | ···················· |
| Q | 3156 hCoV-19/England/QEUH-A61CF0/2020\|EPI_ISL_628647\|20 | ···················· | ···················· | ···················· | ···················· | ···················· | ···················· |
| Q | 3157 hCoV-19/England/QEUH-A61D2A/2020\|EPI_ISL_628648\|20 | ···················· | ···················· | ···················· | ···················· | ···················· | ···················· |
| Q | 3158 hCoV-19/England/QEUH-A61D39/2020\|EPI_ISL_628649\|20 | ···················· | ···················· | ···················· | ···················· | ···················· | ···················· |
| Q | 3159 hCoV-19/England/QEUH-A61D48/2020\|EPI_ISL_628650\|20 | ···················· | ···················· | ···················· | ···················· | ···················· | ···················· |
| Q | 3160 hCoV-19/England/QEUH-A61D84/2020\|EPI_ISL_628651\|20 | ···················· | ···················· | ···················· | ···················· | ···················· | ···················· |
| Q | 3161 hCoV-19/England/QEUH-A61DB1/2020\|EPI_ISL_628652\|20 | ···················· | ···················· | ···················· | ···················· | ···················· | ···················· |
| Q | 3162 hCoV-19/England/QEUH-A61E54/2020\|EPI_ISL_628653\|20 | ···················· | ···················· | ···················· | ···················· | ···················· | ···················· |
| Q | 3163 hCoV-19/England/QEUH-A61E63/2020\|EPI_ISL_628654\|20 | ···················· | ···················· | ···················· | ···················· | ···················· | ···················· |
| Q | 3164 hCoV-19/England/QEUH-A61E72/2020\|EPI_ISL_628655\|20 | ···················· | ·A·················· | ···················· | ···················· | ···················· | ···················· |
| Q | 3165 hCoV-19/England/QEUH-A61E81/2020\|EPI_ISL_628656\|20 | ···················· | ···················· | ···················· | ···················· | ···················· | ···················· |
| Q | 3166 hCoV-19/England/QEUH-A61E90/2020\|EPI_ISL_628657\|20 | ···················· | ···················· | ···················· | ···················· | ···················· | ···················· |
| Q | 3167 hCoV-19/England/QEUH-A61EBE/2020\|EPI_ISL_628658\|20 | ···················· | ···················· | ···················· | ···················· | ···················· | ···················· |
| Q | 3168 hCoV-19/England/QEUH-A63A4F/2020\|EPI_ISL_628659\|20 | ···················· | ···················· | ···················· | ···················· | ···················· | ···················· |
| Q | 3169 hCoV-19/England/QEUH-A63A7C/2020\|EPI_ISL_628660\|20 | ···················· | ·A·················· | ···················· | ···················· | ···················· | ···················· |
| Q | 3170 hCoV-19/England/QEUH-A63A9A/2020\|EPI_ISL_628661\|20 | ···················· | ···················· | ···················· | ···················· | ···················· | ···················· |
| Q | 3171 hCoV-19/Wales/QEUH-A63AA9/2020\|EPI_ISL_628662\|2020 | ···················· | ···················· | ···················· | ···················· | ···················· | ···················· |
| Q | 3172 hCoV-19/England/QEUH-A63AB8/2020\|EPI_ISL_628663\|20 | ···················· | ···················· | ···················· | ···················· | ···················· | ···················· |
| Q | 3173 hCoV-19/England/QEUH-A63BA6/2020\|EPI_ISL_628664\|20 | ···················· | ···················· | ···················· | ···················· | ···················· | ···················· |
| Q | 3174 hCoV-19/England/QEUH-A63BD3/2020\|EPI_ISL_628665\|20 | ···················· | ···················· | ···················· | ···················· | ···················· | ···················· |
| Q | 3175 hCoV-19/England/QEUH-A63BE2/2020\|EPI_ISL_628666\|20 | ···················· | ···················· | ···················· | ···················· | ···················· | ···················· |
| Q | 3176 hCoV-19/England/QEUH-A63C1C/2020\|EPI_ISL_628667\|20 | ···················· | ···················· | ···················· | ···················· | ···················· | ···················· |
| Q | 3177 hCoV-19/England/QEUH-A63C3A/2020\|EPI_ISL_628668\|20 | ···················· | ···················· | ···················· | ···················· | ···················· | ···················· |
| Q | 3178 hCoV-19/England/QEUH-A63D73/2020\|EPI_ISL_628669\|20 | ···················· | ·A·················· | ···················· | ···················· | ···················· | ···················· |
| Q | 3179 hCoV-19/England/QEUH-A63D82/2020\|EPI_ISL_628670\|20 | ···················· | ···················· | ···················· | ···················· | ···················· | ···················· |
| Q | 3180 hCoV-19/England/QEUH-A63D91/2020\|EPI_ISL_628671\|20 | ···················· | ···················· | ···················· | ···················· | ···················· | ···················· |
| Q | 3181 hCoV-19/England/QEUH-A63DA0/2020\|EPI_ISL_628672\|20 | ···················· | ·A·················· | ···················· | ···················· | ···················· | ···················· |
| Q | 3182 hCoV-19/England/QEUH-A63DBF/2020\|EPI_ISL_628673\|20 | ···················· | ···················· | ···················· | ···················· | ···················· | ···················· |
| Q | 3183 hCoV-19/England/QEUH-A63DCE/2020\|EPI_ISL_628674\|20 | ···················· | ···················· | ···················· | ···················· | ···················· | ···················· |
| Q | 3184 hCoV-19/England/QEUH-A63EE9/2020\|EPI_ISL_628675\|20 | ···················· | ···················· | ···················· | ···················· | ···················· | ···················· |
| Q | 3185 hCoV-19/England/QEUH-A63EF8/2020\|EPI_ISL_628676\|20 | ···················· | ···················· | ···················· | ···················· | ···················· | ···················· |
| Q | 3186 hCoV-19/England/QEUH-A63F13/2020\|EPI_ISL_628677\|20 | ···················· | ···················· | ···················· | ···················· | ···················· | ···················· |
| Q | 3187 hCoV-19/England/QEUH-A63F8C/2020\|EPI_ISL_628678\|20 | ···················· | ···················· | ···················· | ···················· | ···················· | ···················· |
| Q | 3188 hCoV-19/England/QEUH-A63FAA/2020\|EPI_ISL_628679\|20 | ···················· | ···················· | ···················· | ···················· | ···················· | ···················· |
| Q | 3189 hCoV-19/England/QEUH-A63FB9/2020\|EPI_ISL_628680\|20 | ···················· | ···················· | ···················· | ···················· | ···················· | ···················· |
| Q | 3190 hCoV-19/England/QEUH-A63FC8/2020\|EPI_ISL_628681\|20 | ···················· | ···················· | ···················· | ···················· | ···················· | ···················· |
| Q | 3191 hCoV-19/England/QEUH-A63FD7/2020\|EPI_ISL_628682\|20 | ···················· | ···················· | ···················· | ···················· | ···················· | ···················· |
| Q | 3192 hCoV-19/England/QEUH-A63FF5/2020\|EPI_ISL_628683\|20 | ···················· | ···················· | ···················· | ···················· | ···················· | ···················· |
| Q | 3193 hCoV-19/England/QEUH-A6401F/2020\|EPI_ISL_628684\|20 | ···················· | ···················· | ···················· | ···················· | ···················· | ···················· |
| Q | 3194 hCoV-19/England/QEUH-A6405B/2020\|EPI_ISL_628685\|20 | ···················· | ···················· | ···················· | ···················· | ···················· | ···················· |
| Q | 3195 hCoV-19/England/QEUH-A640A6/2020\|EPI_ISL_628686\|20 | ···················· | ···················· | ···················· | ···················· | ···················· | ···················· |
| Q | 3196 hCoV-19/England/QEUH-A640D3/2020\|EPI_ISL_628687\|20 | ···················· | ···················· | ···················· | ···················· | ···················· | ···················· |
| Q | 3197 hCoV-19/England/QEUH-A64307/2020\|EPI_ISL_628688\|20 | ···················· | ···················· | ···················· | ···················· | ···················· | ···················· |
| Q | 3198 hCoV-19/England/QEUH-A64334/2020\|EPI_ISL_628689\|20 | ···················· | ···················· | ···················· | ···················· | ···················· | ···················· |
| Q | 3199 hCoV-19/England/QEUH-A64352/2020\|EPI_ISL_628690\|20 | ···················· | ·A·················· | ···················· | ···················· | ···················· | ···A················ |
| Q | 3200 hCoV-19/England/QEUH-A64361/2020\|EPI_ISL_628691\|20 | ···················· | ···················· | ···················· | ···················· | ···················· | ···················· |
| Q | 3201 hCoV-19/England/QEUH-A6438F/2020\|EPI_ISL_628692\|20 | ···················· | ···················· | ···················· | ···················· | ···················· | ···················· |
| Q | 3202 hCoV-19/England/QEUH-A643BC/2020\|EPI_ISL_628693\|20 | ···················· | ···················· | ···················· | ···················· | ···················· | ···················· |
| Q | 3203 hCoV-19/England/QEUH-A643CB/2020\|EPI_ISL_628694\|20 | ···················· | ···················· | ···················· | ···················· | ···················· | ···················· |
| Q | 3204 hCoV-19/England/QEUH-A643E9/2020\|EPI_ISL_628695\|20 | ···················· | ···················· | ···················· | ···················· | ···················· | ···················· |
| Q | 3205 hCoV-19/England/QEUH-A643F8/2020\|EPI_ISL_628696\|20 | ···················· | ···················· | ···················· | ···················· | ···················· | ···················· |
| Q | 3206 hCoV-19/England/QEUH-A64422/2020\|EPI_ISL_628697\|20 | ···················· | ···················· | ···················· | ···················· | ···················· | ···················· |
| Q | 3207 hCoV-19/England/QEUH-A64431/2020\|EPI_ISL_628698\|20 | ···················· | ···················· | ···················· | ···················· | ···················· | ···················· |
| Q | 3208 hCoV-19/England/QEUH-A64440/2020\|EPI_ISL_628699\|20 | ···················· | ···················· | ···················· | ···················· | ···················· | ···················· |
| Q | 3209 hCoV-19/England/QEUH-AD29A8/2020\|EPI_ISL_629906\|20 | ···················· | ···················· | ···················· | ···················· | ···················· | ···················· |
| Q | 3210 hCoV-19/England/QEUH-AD2AE1/2020\|EPI_ISL_629908\|20 | ···················· | ···················· | ···················· | ···················· | ···················· | ···················· |
| Q | 3211 hCoV-19/England/QEUH-AD2E21/2020\|EPI_ISL_629910\|20 | ···················· | ·A·················· | ···················· | ···················· | ···················· | ···A················ |
| Q | 3212 hCoV-19/England/QEUH-AD254A/2020\|EPI_ISL_629911\|20 | ···················· | ···················· | ···················· | ···················· | ···················· | ···················· |
| Q | 3213 hCoV-19/England/QEUH-AD04A9/2020\|EPI_ISL_629913\|20 | ···················· | ···················· | ···················· | ···················· | ······T············· | ···················· |

| | | | | | | | | |
|---|---|---|---|---|---|---|---|---|
| Q | 3214 | hCoV-19/England/QEUH-AD2DCA/2020\|EPI_ISL_629915\|20 | ···················· | ···················· | ····················· | ···················· | ···················· | ···················· |
| Q | 3215 | hCoV-19/England/QEUH-AD2322/2020\|EPI_ISL_629916\|20 | ···················· | ···················· | ····················· | ···················· | ···················· | ···················· |
| Q | 3216 | hCoV-19/England/QEUH-AD1D25/2020\|EPI_ISL_629917\|20 | ···················· | ···················· | ····················· | ···················· | ···················· | ···················· |
| Q | 3217 | hCoV-19/England/QEUH-AD22BC/2020\|EPI_ISL_629918\|20 | ···················· | ···················· | ····················· | ···················· | ···················· | ···················· |
| Q | 3218 | hCoV-19/England/QEUH-AD3A95/2020\|EPI_ISL_629919\|20 | ···················· | ···················· | ····················· | ···················· | ···················· | ···················· |
| Q | 3219 | hCoV-19/England/QEUH-AD06EF/2020\|EPI_ISL_629920\|20 | ···················· | ···················· | ····················· | ···················· | ···················· | ···················· |
| Q | 3220 | hCoV-19/England/QEUH-AD1CCE/2020\|EPI_ISL_629921\|20 | ···················· | ···················· | ····················· | ···················· | ···················· | ···················· |
| Q | 3221 | hCoV-19/England/QEUH-AD2D42/2020\|EPI_ISL_629922\|20 | ···················· | ···················· | ····················· | ···················· | ···················· | ···················· |
| Q | 3222 | hCoV-19/England/QEUH-AD2313/2020\|EPI_ISL_629923\|20 | ···················· | ···················· | ····················· | ···················· | ···················· | ···················· |
| Q | 3223 | hCoV-19/England/QEUH-AD0913/2020\|EPI_ISL_629924\|20 | ···················· | ···················· | ····················· | ···················· | ···················· | ···················· |
| Q | 3224 | hCoV-19/England/QEUH-AD08F8/2020\|EPI_ISL_629925\|20 | ···················· | ···················· | ····················· | ···················· | ···················· | ···················· |
| Q | 3225 | hCoV-19/England/QEUH-AD0324/2020\|EPI_ISL_629926\|20 | ···················· | ···················· | ····················· | ···················· | ···················· | ···················· |
| Q | 3226 | hCoV-19/England/QEUH-AD3A77/2020\|EPI_ISL_629927\|20 | ···················· | ···················· | ····················· | ···················· | ···················· | ···················· |
| Q | 3227 | hCoV-19/England/QEUH-AD3D6F/2020\|EPI_ISL_629971\|20 | ···················· | ···················· | ····················· | ···················· | ···················· | ···················· |
| Q | 3228 | hCoV-19/England/QEUH-AD2AA5/2020\|EPI_ISL_629972\|20 | ···················· | ·A·················· | ····················· | ···················· | ···················· | ··A················· |
| Q | 3229 | hCoV-19/England/QEUH-AD3AE0/2020\|EPI_ISL_629973\|20 | ···················· | ···················· | ····················· | ···················· | ···················· | ···················· |
| Q | 3230 | hCoV-19/England/QEUH-AD17FA/2020\|EPI_ISL_629975\|20 | ···················· | ···················· | ····················· | ···················· | ···················· | ···················· |
| Q | 3231 | hCoV-19/England/QEUH-AD2753/2020\|EPI_ISL_629976\|20 | ···················· | ···················· | ····················· | ···················· | ···················· | ···················· |
| Q | 3232 | hCoV-19/England/QEUH-AD061C/2020\|EPI_ISL_629979\|20 | ···················· | ·A·················· | ····················· | ···················· | ···················· | ···················· |
| Q | 3233 | hCoV-19/England/QEUH-AD2D51/2020\|EPI_ISL_629980\|20 | ···················· | ···················· | ····················· | ···················· | ···················· | ···················· |
| Q | 3234 | hCoV-19/England/QEUH-AD2629/2020\|EPI_ISL_629981\|20 | ···················· | ···················· | ····················· | ···················· | ···················· | ···················· |
| Q | 3235 | hCoV-19/England/QEUH-AD1F7A/2020\|EPI_ISL_629982\|20 | ···················· | ···················· | ····················· | ···················· | ···················· | ···················· |
| Q | 3236 | hCoV-19/Wales/QEUH-AD3E20/2020\|EPI_ISL_629983\|2020 | ···················· | ···················· | ····················· | ···················· | ···················· | ···················· |
| Q | 3237 | hCoV-19/England/QEUH-AD044F/2020\|EPI_ISL_629984\|20 | ···················· | ···················· | ····················· | ···················· | ···················· | ···················· |
| Q | 3238 | hCoV-19/England/QEUH-AD17AF/2020\|EPI_ISL_629985\|20 | ···················· | ···················· | ····················· | ···················· | ···················· | ···················· |
| Q | 3239 | hCoV-19/England/QEUH-AD06C1/2020\|EPI_ISL_629986\|20 | ···················· | ···················· | ····················· | ···················· | ···················· | ···················· |
| Q | 3240 | hCoV-19/England/QEUH-AD29B7/2020\|EPI_ISL_629988\|20 | ···················· | ···················· | ····················· | ···················· | ···················· | ···················· |
| Q | 3241 | hCoV-19/England/QEUH-AD25E0/2020\|EPI_ISL_629989\|20 | ···················· | ···················· | ····················· | ···················· | ···················· | ···················· |
| Q | 3242 | hCoV-19/England/QEUH-AD3E11/2020\|EPI_ISL_629990\|20 | ···················· | ···················· | ····················· | ···················· | ···················· | ···················· |
| Q | 3243 | hCoV-19/England/QEUH-AD346A/2020\|EPI_ISL_629991\|20 | ···················· | ···················· | ····················· | ···················· | ···················· | ···················· |
| Q | 3244 | hCoV-19/England/QEUH-AD1C82/2020\|EPI_ISL_629992\|20 | ···················· | ···················· | ····················· | ···················· | ···················· | ···················· |
| Q | 3245 | hCoV-19/England/QEUH-AD3A2C/2020\|EPI_ISL_629993\|20 | ···················· | ···················· | ····················· | ···················· | ···················· | ···················· |
| Q | 3246 | hCoV-19/England/QEUH-AD09D7/2020\|EPI_ISL_629995\|20 | ···················· | ···················· | ····················· | ···················· | ···················· | ···················· |
| Q | 3247 | hCoV-19/England/QEUH-AD1D34/2020\|EPI_ISL_629996\|20 | ···················· | ···················· | ····················· | ···················· | ···················· | ···················· |
| Q | 3248 | hCoV-19/England/QEUH-AD17DC/2020\|EPI_ISL_629997\|20 | ···················· | ···················· | ····················· | ···················· | ···················· | ···················· |
| Q | 3249 | hCoV-19/England/QEUH-AD2A78/2020\|EPI_ISL_629998\|20 | ···················· | ···················· | ····················· | ···················· | ···················· | ···················· |
| Q | 3250 | hCoV-19/England/QEUH-AD24D4/2020\|EPI_ISL_629999\|20 | ···················· | ···················· | ····················· | ···················· | ···················· | ···················· |
| Q | 3251 | hCoV-19/England/QEUH-AAE7E7/2020\|EPI_ISL_630900\|20 | ···················· | ···················· | ····················· | ···················· | ···················· | ···················· |
| Q | 3252 | hCoV-19/England/ALDP-AA4D1C/2020\|EPI_ISL_630901\|20 | ···················· | ···················· | ····················· | ···················· | ···················· | ···················· |
| Q | 3253 | hCoV-19/England/ALDP-AA4F8F/2020\|EPI_ISL_630902\|20 | ···················· | ···················· | ····················· | ···················· | ···················· | ···················· |
| Q | 3254 | hCoV-19/England/ALDP-AA4D3A/2020\|EPI_ISL_630904\|20 | ···················· | ···················· | ····················· | ···················· | ···················· | ···················· |
| Q | 3255 | hCoV-19/England/ALDP-AA50E5/2020\|EPI_ISL_630905\|20 | ···················· | ···················· | ····················· | ···················· | ···················· | ···················· |
| Q | 3256 | hCoV-19/England/ALDP-AA1B7F/2020\|EPI_ISL_630906\|20 | ···················· | ···················· | ····················· | ···················· | ···················· | ···················· |
| Q | 3257 | hCoV-19/England/ALDP-AA2613/2020\|EPI_ISL_630907\|20 | ···················· | ···················· | ····················· | ···················· | ···················· | ···················· |
| Q | 3258 | hCoV-19/England/ALDP-AA3919/2020\|EPI_ISL_630908\|20 | ···················· | ···················· | ····················· | ···················· | ···················· | ···················· |
| Q | 3259 | hCoV-19/England/ALDP-AA306F/2020\|EPI_ISL_630909\|20 | ···················· | ···················· | ····················· | ···················· | ···················· | ···················· |
| Q | 3260 | hCoV-19/England/ALDP-AA4E73/2020\|EPI_ISL_630952\|20 | ···················· | ·········T·········· | ····················· | ···················· | ···················· | ···················· |
| Q | 3261 | hCoV-19/England/ALDP-AA390A/2020\|EPI_ISL_630954\|20 | ···················· | ···················· | ····················· | ···················· | ···················· | ···················· |
| Q | 3262 | hCoV-19/England/QEUH-AAE064/2020\|EPI_ISL_630955\|20 | ···················· | ···················· | ····················· | ···················· | ···················· | ···················· |
| Q | 3263 | hCoV-19/England/QEUH-AAE055/2020\|EPI_ISL_630959\|20 | ···················· | ···················· | ····················· | ···················· | ···················· | ···················· |
| Q | 3264 | hCoV-19/England/ALDP-AA5249/2020\|EPI_ISL_630960\|20 | ···················· | ···················· | ····················· | ···················· | ···················· | ···················· |
| Q | 3265 | hCoV-19/England/ALDP-AA317B/2020\|EPI_ISL_630961\|20 | ···················· | ···················· | ····················· | ···················· | ···················· | ···················· |
| Q | 3266 | hCoV-19/England/ALDP-AA2FFA/2020\|EPI_ISL_630962\|20 | ···················· | ···················· | ····················· | ···················· | ···················· | ···················· |
| Q | 3267 | hCoV-19/England/ALDP-AA4B30/2020\|EPI_ISL_630963\|20 | ···················· | ···················· | ····················· | ···················· | ···················· | ···················· |
| Q | 3268 | hCoV-19/England/ALDP-AA509A/2020\|EPI_ISL_630964\|20 | ···················· | ···················· | ····················· | ···················· | ···················· | ···················· |
| Q | 3269 | hCoV-19/England/QEUH-AAF31F/2020\|EPI_ISL_630965\|20 | ···················· | ·A·················· | ····················· | ···················· | ···················· | ···················· |
| Q | 3270 | hCoV-19/England/ALDP-AA4C79/2020\|EPI_ISL_630966\|20 | ···················· | ···················· | ····················· | ···················· | ···················· | ···················· |
| Q | 3271 | hCoV-19/England/ALDP-AA26F5/2020\|EPI_ISL_630968\|20 | ···················· | ···················· | ····················· | ···················· | ···················· | ···················· |
| Q | 3272 | hCoV-19/England/ALDP-AA3955/2020\|EPI_ISL_630969\|20 | ···················· | ···················· | ····················· | ···················· | ···················· | ···················· |
| Q | 3273 | hCoV-19/England/ALDP-AA25E9/2020\|EPI_ISL_630970\|20 | ···················· | ···················· | ····················· | ···················· | ···················· | ···················· |
| Q | 3274 | hCoV-19/England/ALDP-ABD211/2020\|EPI_ISL_630971\|20 | ···················· | ···················· | ····················· | ···················· | ···················· | ···················· |
| Q | 3275 | hCoV-19/England/CAMC-AAF634/2020\|EPI_ISL_630972\|20 | ···················· | ···················· | ····················· | ···················· | ···················· | ···················· |
| Q | 3276 | hCoV-19/England/MILK-ABC1D9/2020\|EPI_ISL_630973\|20 | ···················· | ···················· | ····················· | ···················· | ···················· | ···················· |
| Q | 3277 | hCoV-19/England/MILK-ABC50A/2020\|EPI_ISL_630974\|20 | ···················· | ···················· | ····················· | ···················· | ···················· | ···················· |
| Q | 3278 | hCoV-19/England/ALDP-AC12FC/2020\|EPI_ISL_630975\|20 | ···················· | ···················· | ····················· | ···················· | ···················· | ···················· |
| Q | 3279 | hCoV-19/England/ALDP-ABD31E/2020\|EPI_ISL_630976\|20 | ···················· | ···················· | ····················· | ···················· | ···················· | ···················· |
| Q | 3280 | hCoV-19/England/CAMC-AAF5BF/2020\|EPI_ISL_630977\|20 | ···················· | ···················· | ····················· | ···················· | ···················· | ···················· |
| Q | 3281 | hCoV-19/England/MILK-ABBFEF/2020\|EPI_ISL_630978\|20 | ···················· | ···················· | ····················· | ···················· | ···················· | ···················· |
| Q | 3282 | hCoV-19/England/CAMC-AAF625/2020\|EPI_ISL_630979\|20 | ···················· | ···················· | ····················· | ···················· | ···················· | ···················· |
| Q | 3283 | hCoV-19/England/MILK-ABC0EB/2020\|EPI_ISL_630981\|20 | ···················· | ···················· | ····················· | ···················· | ···················· | ···················· |
| Q | 3284 | hCoV-19/England/ALDP-AC11FF/2020\|EPI_ISL_630982\|20 | ···················· | ···················· | ····················· | ···················· | ···················· | ···················· |
| Q | 3285 | hCoV-19/England/ALDP-AC5D8E/2020\|EPI_ISL_630983\|20 | ···················· | ···················· | ····················· | ···················· | ···················· | ···················· |
| Q | 3286 | hCoV-19/England/CAMC-AAF7E6/2020\|EPI_ISL_630984\|20 | ···················· | ···················· | ····················· | ···················· | ···················· | ···················· |
| Q | 3287 | hCoV-19/England/ALDP-AC11C2/2020\|EPI_ISL_630985\|20 | ···················· | ···················· | ····················· | ···················· | ···················· | ···················· |
| Q | 3288 | hCoV-19/England/MILK-ABBB81/2020\|EPI_ISL_630986\|20 | ···················· | ···················· | ····················· | ···················· | ···················· | ···················· |
| Q | 3289 | hCoV-19/England/CAMC-AAFA38/2020\|EPI_ISL_630988\|20 | ···················· | ···················· | ····················· | ···················· | ···················· | ···················· |
| Q | 3290 | hCoV-19/England/MILK-ABBF0D/2020\|EPI_ISL_630990\|20 | ···················· | ···················· | ····················· | ···················· | ···················· | ···················· |
| Q | 3291 | hCoV-19/England/MILK-ABB5CF/2020\|EPI_ISL_630991\|20 | ···················· | ···················· | ····················· | ···················· | ···················· | ···················· |
| Q | 3292 | hCoV-19/England/MILK-ABC090/2020\|EPI_ISL_630993\|20 | ···················· | ···················· | ····················· | ···················· | ···················· | ···················· |
| Q | 3293 | hCoV-19/England/ALDP-ABD32D/2020\|EPI_ISL_630994\|20 | ···················· | ···················· | ····················· | ···················· | ···················· | ···················· |

| | | | | | | | |
|---|---|---|---|---|---|---|---|
| Q | 3294 hCoV-19/England/MILK-ABC27C/2020\|EPI_ISL_630995\|20 | ···················· | ···················· | ···················· | ···················· | ···················· | ···················· |
| Q | 3295 hCoV-19/England/CAMC-AAF93B/2020\|EPI_ISL_630996\|20 | ···················· | ···················· | ···················· | ···················· | ···················· | ···················· |
| Q | 3296 hCoV-19/England/CAMC-AAF4EF/2020\|EPI_ISL_630997\|20 | ···················· | ···················· | ···················· | ···················· | ···················· | ···················· |
| Q | 3297 hCoV-19/England/MILK-ABBFD0/2020\|EPI_ISL_630998\|20 | ···················· | ···················· | ···················· | ···················· | ···················· | ···················· |
| Q | 3298 hCoV-19/England/ALDP-AC5E03/2020\|EPI_ISL_630999\|20 | ···················· | ···················· | ···················· | ···················· | ···················· | ···················· |
| Q | 3299 hCoV-19/England/QEUH-AE4F1A/2020\|EPI_ISL_633154\|20 | ···················· | ···················· | ···················· | ···················· | ···················· | ···················· |
| Q | 3300 hCoV-19/England/CAMC-AEA758/2020\|EPI_ISL_633155\|20 | ···················· | ···················· | ···················· | ···················· | ···················· | ···················· |
| Q | 3301 hCoV-19/England/CAMC-AEAE62/2020\|EPI_ISL_633156\|20 | ···················· | ···················· | ···················· | ···················· | ···················· | ···················· |
| Q | 3302 hCoV-19/England/CAMC-AEADB0/2020\|EPI_ISL_633157\|20 | ···················· | ···················· | ···················· | ···················· | ·····T·············· | ···················· |
| Q | 3303 hCoV-19/England/QEUH-AE4B16/2020\|EPI_ISL_633158\|20 | ···················· | ···················· | ···················· | ···················· | ···················· | ···················· |
| Q | 3304 hCoV-19/England/CAMC-AEB001/2020\|EPI_ISL_633159\|20 | ···················· | ···················· | ···················· | ···················· | ·····T·············· | ···················· |
| Q | 3305 hCoV-19/England/CAMC-AEABD4/2020\|EPI_ISL_633170\|20 | ···················· | ···················· | ···················· | ···················· | ·····T·············· | ···················· |
| Q | 3306 hCoV-19/England/QEUH-AE4B61/2020\|EPI_ISL_633171\|20 | ···················· | ···················· | ···················· | ···················· | ···················· | ···················· |
| Q | 3307 hCoV-19/England/CAMC-AEB168/2020\|EPI_ISL_633173\|20 | ···················· | ···················· | ···················· | ···················· | ···················· | ···················· |
| Q | 3308 hCoV-19/England/QEUH-AE50AD/2020\|EPI_ISL_633174\|20 | ···················· | ···················· | ···················· | ···················· | ···················· | ···················· |
| Q | 3309 hCoV-19/England/QEUH-AE4CF5/2020\|EPI_ISL_633175\|20 | ···················· | ···················· | ···················· | ···················· | ···················· | ···················· |
| Q | 3310 hCoV-19/England/CAMC-AEAEAE/2020\|EPI_ISL_633176\|20 | ···················· | ···················· | ···················· | ···················· | ···················· | ···················· |
| Q | 3311 hCoV-19/England/CAMC-AEA63D/2020\|EPI_ISL_633177\|20 | ···················· | ···················· | ···················· | ···················· | ···················· | ···················· |
| Q | 3312 hCoV-19/England/QEUH-AE50E9/2020\|EPI_ISL_633178\|20 | ···················· | ···················· | ···················· | ···················· | ···················· | ···················· |
| Q | 3313 hCoV-19/England/QEUH-AE5438/2020\|EPI_ISL_633179\|20 | ···················· | ···················· | ···················· | ···················· | ···················· | ···················· |
| Q | 3314 hCoV-19/England/QEUH-AE4D01/2020\|EPI_ISL_633180\|20 | ···················· | ···················· | ···················· | ···················· | ···················· | ···················· |
| Q | 3315 hCoV-19/England/QEUH-AE4F92/2020\|EPI_ISL_633181\|20 | ···················· | ···················· | ···················· | ···················· | ···················· | ···················· |
| Q | 3316 hCoV-19/England/QEUH-AE5201/2020\|EPI_ISL_633182\|20 | ···················· | ···················· | ···················· | ···················· | ···················· | ···················· |
| Q | 3317 hCoV-19/England/CAMC-AEA6D3/2020\|EPI_ISL_633183\|20 | ···················· | ···················· | ···················· | ···················· | ···················· | ···················· |
| Q | 3318 hCoV-19/England/QEUH-AE52C5/2020\|EPI_ISL_633184\|20 | ···················· | ···················· | ···················· | ···················· | ···················· | ···················· |
| Q | 3319 hCoV-19/England/QEUH-AE49FE/2020\|EPI_ISL_633185\|20 | ···················· | ···················· | ···················· | ···················· | ···················· | ···················· |
| Q | 3320 hCoV-19/England/CAMC-AEA925/2020\|EPI_ISL_633186\|20 | ···················· | ···················· | ···················· | ···················· | ···················· | ···················· |
| Q | 3321 hCoV-19/England/CAMC-AEABC5/2020\|EPI_ISL_633187\|20 | ···················· | ···················· | ···················· | ···················· | ···················· | ···················· |
| Q | 3322 hCoV-19/England/CAMC-AEB1D1/2020\|EPI_ISL_633188\|20 | ···················· | ···················· | ···················· | ···················· | ···················· | ···················· |
| Q | 3323 hCoV-19/England/CAMC-AEAB7A/2020\|EPI_ISL_633189\|20 | ···················· | ···················· | ···················· | ···················· | ···················· | ···················· |
| Q | 3324 hCoV-19/England/MILK-ACC4CE/2020\|EPI_ISL_634481\|20 | ···················· | ···················· | ···················· | ···················· | ···················· | ···················· |
| Q | 3325 hCoV-19/England/MILK-AD011B/2020\|EPI_ISL_634486\|20 | ···················· | ···················· | ···················· | ···················· | ·····T·············· | ···················· |
| Q | 3326 hCoV-19/England/MILK-ACC938/2020\|EPI_ISL_634487\|20 | ···················· | ···················· | ···················· | ···················· | ···················· | ···················· |
| Q | 3327 hCoV-19/England/MILK-ACC974/2020\|EPI_ISL_634488\|20 | ···················· | ···················· | ···················· | ···················· | ···················· | ···················· |
| Q | 3328 hCoV-19/England/MILK-ACC859/2020\|EPI_ISL_634489\|20 | ···················· | ·A·················· | ···················· | ···················· | ···················· | ··A················· |
| Q | 3329 hCoV-19/England/QEUH-A6163E/2020\|EPI_ISL_627292\|20 | ···················· | ···················· | ···················· | ···················· | ···················· | ···················· |
| Q | 3331 hCoV-19/Wales/PHWC-483229/2020\|EPI_ISL_627296\|2020 | ···················· | ···················· | ···················· | ···················· | ···················· | ···················· |
| Q | 3332 hCoV-19/Wales/CVR4740/2020\|EPI_ISL_627332\|2020-09- | ···················· | ···················· | ···················· | ···················· | ···················· | ···················· |
| Q | 3333 hCoV-19/England/QEUH-A4BB0E/2020\|EPI_ISL_628509\|20 | ···················· | ···················· | ···················· | ···················· | ···················· | ···················· |
| Q | 3334 hCoV-19/Scotland/QEUH-A4BBD1/2020\|EPI_ISL_628514\|2 | ···················· | ···················· | ···················· | ···················· | ···················· | ···················· |
| Q | 3335 hCoV-19/England/QEUH-AD2207/2020\|EPI_ISL_629808\|20 | ···················· | ···················· | ···················· | ···················· | ···················· | ···················· |
| Q | 3336 hCoV-19/England/QEUH-AD0931/2020\|EPI_ISL_629809\|20 | ···················· | ···················· | ···················· | ···················· | ···················· | ···················· |
| Q | 3337 hCoV-19/England/QEUH-AD39A7/2020\|EPI_ISL_629810\|20 | ···················· | ···················· | ···················· | ···················· | ···················· | ···················· |
| Q | 3338 hCoV-19/England/QEUH-AD0333/2020\|EPI_ISL_629811\|20 | ···················· | ···················· | ···················· | ···················· | ···················· | ···················· |
| Q | 3339 hCoV-19/England/QEUH-AD26DE/2020\|EPI_ISL_629812\|20 | ···················· | ·A·················· | ···················· | ···················· | ···················· | ··A················· |
| Q | 3340 hCoV-19/England/QEUH-AD09AA/2020\|EPI_ISL_629813\|20 | ···················· | ···················· | ···················· | ···················· | ···················· | ···················· |
| Q | 3341 hCoV-19/England/QEUH-AD2DAC/2020\|EPI_ISL_629815\|20 | ···················· | ···················· | ···················· | ···················· | ···················· | ···················· |
| Q | 3342 hCoV-19/England/QEUH-AD0E8D/2020\|EPI_ISL_629816\|20 | ···················· | ···················· | ···················· | ···················· | ···················· | ···················· |
| Q | 3343 hCoV-19/England/QEUH-AD2F97/2020\|EPI_ISL_629817\|20 | ···················· | ···················· | ···················· | ···················· | ···················· | ···················· |
| Q | 3344 hCoV-19/England/QEUH-AD2647/2020\|EPI_ISL_629818\|20 | ···················· | ···················· | ···················· | ···················· | ···················· | ···················· |
| Q | 3345 hCoV-19/England/QEUH-AD22E9/2020\|EPI_ISL_629820\|20 | ···················· | ···················· | ···················· | ···················· | ···················· | ···················· |
| Q | 3346 hCoV-19/England/QEUH-AD06FE/2020\|EPI_ISL_629821\|20 | ···················· | ···················· | ···················· | ···················· | ···················· | ···················· |
| Q | 3347 hCoV-19/England/QEUH-AD0272/2020\|EPI_ISL_629822\|20 | ···················· | ···················· | ···················· | ···················· | ···················· | ···················· |
| Q | 3348 hCoV-19/England/QEUH-AD1EB9/2020\|EPI_ISL_629824\|20 | ···················· | ···················· | ···················· | ···················· | ···················· | ···················· |
| Q | 3349 hCoV-19/England/QEUH-AD2F6A/2020\|EPI_ISL_629825\|20 | ···················· | ···················· | ···················· | ···················· | ···················· | ···················· |
| Q | 3350 hCoV-19/England/QEUH-AD0649/2020\|EPI_ISL_629826\|20 | ···················· | ···················· | ···················· | ···················· | ···················· | ···················· |
| Q | 3351 hCoV-19/England/QEUH-AD39E3/2020\|EPI_ISL_629827\|20 | ···················· | ···················· | ···················· | ···················· | ···················· | ···················· |
| Q | 3352 hCoV-19/England/QEUH-AD2E9A/2020\|EPI_ISL_629828\|20 | ···················· | ···················· | ···················· | ···················· | ···················· | ···················· |
| Q | 3353 hCoV-19/England/QEUH-AD1824/2020\|EPI_ISL_629845\|20 | ···················· | ···················· | ···················· | ···················· | ···················· | ···················· |
| Q | 3354 hCoV-19/England/QEUH-AD633D/2020\|EPI_ISL_629846\|20 | ···················· | ···················· | ···················· | ···················· | ···················· | ···················· |
| Q | 3355 hCoV-19/England/ALDP-AA4031/2020\|EPI_ISL_630800\|20 | ···················· | ···················· | ···················· | ···················· | ···················· | ···················· |
| Q | 3356 hCoV-19/England/ALDP-AA31B7/2020\|EPI_ISL_630801\|20 | ···················· | ···················· | ···················· | ···················· | ···················· | ···················· |
| Q | 3357 hCoV-19/England/ALDP-AA25CB/2020\|EPI_ISL_630802\|20 | ···················· | ···················· | ···················· | ···················· | ···················· | ···················· |
| Q | 3358 hCoV-19/England/ALDP-AA4B03/2020\|EPI_ISL_630803\|20 | ···················· | ···················· | ···················· | ···················· | ···················· | ···················· |
| Q | 3359 hCoV-19/England/QEUH-AAE116/2020\|EPI_ISL_630804\|20 | ···················· | ···················· | ···················· | ···················· | ···················· | ···················· |
| Q | 3360 hCoV-19/England/ALDP-AA1BF7/2020\|EPI_ISL_630805\|20 | ···················· | ···················· | ···················· | ···················· | ···················· | ···················· |
| Q | 3361 hCoV-19/England/ALDP-AA512E/2020\|EPI_ISL_630807\|20 | ···················· | ···················· | ···················· | ···················· | ···················· | ···················· |
| Q | 3362 hCoV-19/England/ALDP-AA4FE9/2020\|EPI_ISL_630808\|20 | ···················· | ···················· | ···················· | ···················· | ···················· | ···················· |
| Q | 3363 hCoV-19/England/ALDP-AA3FEA/2020\|EPI_ISL_630809\|20 | ···················· | ···················· | ···················· | ···················· | ···················· | ···················· |
| Q | 3364 hCoV-19/England/ALDP-AA2A9F/2020\|EPI_ISL_630823\|20 | ···················· | ···················· | ···················· | ···················· | ···················· | ···················· |
| Q | 3365 hCoV-19/England/QEUH-AAE76F/2020\|EPI_ISL_630824\|20 | ···················· | ···················· | ···················· | ···················· | ···················· | ···················· |
| Q | 3366 hCoV-19/USA/NY-NYCPHL-001070/2020\|EPI_ISL_633071\|2 | ···················· | ···················· | ···················· | ···················· | ···················· | ···················· |
| Q | 3367 hCoV-19/USA/NY-NYCPHL-001071/2020\|EPI_ISL_633072\|2 | ···················· | ···················· | ···················· | ···················· | ···················· | ···················· |
| Q | 3368 hCoV-19/USA/NY-NYCPHL-001072/2020\|EPI_ISL_633073\|2 | ···················· | ···················· | ···················· | ···················· | ···················· | ···················· |
| Q | 3369 hCoV-19/USA/NY-NYCPHL-001073/2020\|EPI_ISL_633074\|2 | ···················· | ···················· | ···················· | ···················· | ···················· | ···················· |
| Q | 3370 hCoV-19/USA/NY-NYCPHL-001074/2020\|EPI_ISL_633075\|2 | ···················· | ···················· | ···················· | ···················· | ···················· | ···················· |
| Q | 3371 hCoV-19/USA/NY-NYCPHL-001075/2020\|EPI_ISL_633076\|2 | ···················· | ···················· | ···················· | ···················· | ···················· | ···················· |
| Q | 3372 hCoV-19/USA/NY-NYCPHL-001077/2020\|EPI_ISL_633078\|2 | ···················· | ···················· | ···················· | ···················· | ···················· | ···················· |
| Q | 3373 hCoV-19/USA/NY-NYCPHL-001078/2020\|EPI_ISL_633079\|2 | ···················· | ···················· | ···················· | ···················· | ···················· | ···················· |
| Q | 3374 hCoV-19/USA/NY-NYCPHL-001079/2020\|EPI_ISL_633080\|2 | ···················· | ···················· | ···················· | ···················· | ···················· | ···················· |

| | | | | | | | |
|---|---|---|---|---|---|---|---|
| Q | 3375 hCoV-19/USA/NY-NYCPHL-001080/2020\|EPI_ISL_633081\|2 | ···················· | ···················· | ···················· | ···················· | ···················· | ···················· |
| Q | 3376 hCoV-19/USA/NY-NYCPHL-001081/2020\|EPI_ISL_633082\|2 | ···················· | ·A·················· | ···················· | ···················· | ···················· | ···················· |
| Q | 3377 hCoV-19/USA/MD-MDH-0301/2020\|EPI_ISL_633083\|2020-0 | ···················· | ···················· | ···················· | ···················· | ···················· | ···················· |
| Q | 3378 hCoV-19/USA/MD-MDH-0302/2020\|EPI_ISL_633084\|2020-1 | ···················· | ···················· | ···················· | ···················· | ···················· | ···················· |
| Q | 3379 hCoV-19/USA/MD-MDH-0303/2020\|EPI_ISL_633085\|2020-1 | ···················· | ···················· | ···················· | ···················· | ···················· | ···················· |
| Q | 3380 hCoV-19/USA/MD-MDH-0305/2020\|EPI_ISL_633086\|2020-1 | ···················· | ···················· | ···················· | ···················· | ···················· | ···················· |
| Q | 3381 hCoV-19/USA/MD-MDH-0306/2020\|EPI_ISL_633087\|2020-1 | ···················· | ···················· | ···················· | ···················· | ···················· | ···················· |
| Q | 3382 hCoV-19/USA/MD-MDH-0309/2020\|EPI_ISL_633088\|2020-1 | ···················· | ···················· | ···················· | ···················· | ···················· | ···················· |
| Q | 3383 hCoV-19/USA/MD-MDH-0313/2020\|EPI_ISL_633089\|2020-1 | ···················· | ···················· | ···················· | ···················· | ···················· | ···················· |
| Q | 3384 hCoV-19/England/CAMC-AEB14A/2020\|EPI_ISL_633100\|20 | ···················· | ···················· | ···················· | ···················· | ···················· | ···················· |
| Q | 3385 hCoV-19/England/CAMC-AEAFBA/2020\|EPI_ISL_633101\|20 | ···················· | ···················· | ···················· | ···················· | ···················· | ···················· |
| Q | 3386 hCoV-19/England/CAMC-AEA952/2020\|EPI_ISL_633102\|20 | ···················· | ···················· | ···················· | ···················· | ······T············· | ···················· |
| Q | 3387 hCoV-19/England/QEUH-AE4E59/2020\|EPI_ISL_633103\|20 | ···················· | ···················· | ···················· | ···················· | ···················· | ···················· |
| Q | 3388 hCoV-19/England/CAMC-AEB1B3/2020\|EPI_ISL_633104\|20 | ···················· | ···················· | ···················· | ···················· | ···················· | ···················· |
| Q | 3389 hCoV-19/England/CAMC-AEB0E3/2020\|EPI_ISL_633105\|20 | ···················· | ···················· | ···················· | ···················· | ···················· | ···················· |
| Q | 3390 hCoV-19/England/CAMC-AEACC2/2020\|EPI_ISL_633106\|20 | ···················· | ···················· | ···················· | ···················· | ···················· | ···················· |
| Q | 3391 hCoV-19/England/CAMC-AEAEDB/2020\|EPI_ISL_633107\|20 | ···················· | ···················· | ···················· | ···················· | ···················· | ···················· |
| Q | 3392 hCoV-19/England/CAMC-AEA855/2020\|EPI_ISL_633109\|20 | ········Y··········· | ···················· | ···················· | ···················· | ···················· | ···················· |
| Q | 3393 hCoV-19/England/CAMC-AEAAD7/2020\|EPI_ISL_633110\|20 | ···················· | ···················· | ···················· | ···················· | ···················· | ···················· |
| Q | 3394 hCoV-19/England/QEUH-AE4B52/2020\|EPI_ISL_633111\|20 | ···················· | ···················· | ···················· | ···················· | ···················· | ···················· |
| Q | 3395 hCoV-19/England/QEUH-AE4DA7/2020\|EPI_ISL_633112\|20 | ···················· | ···················· | ···················· | ···················· | ···················· | ···················· |
| Q | 3396 hCoV-19/England/QEUH-AE4FB0/2020\|EPI_ISL_633113\|20 | ···················· | ···················· | ···················· | ···················· | ···················· | ···················· |
| Q | 3397 hCoV-19/England/QEUH-AE534A/2020\|EPI_ISL_633115\|20 | ···················· | ···················· | ···················· | ···················· | ······T············· | ···················· |
| Q | 3398 hCoV-19/England/CAMC-AEAB98/2020\|EPI_ISL_633116\|20 | ···················· | ···················· | ···················· | ···················· | ···················· | ···················· |
| Q | 3399 hCoV-19/England/CAMC-AEAF8D/2020\|EPI_ISL_633117\|20 | ···················· | ···················· | ···················· | ···················· | ···················· | ···················· |
| Q | 3400 hCoV-19/England/QEUH-AE5122/2020\|EPI_ISL_633119\|20 | ···················· | ····A··············· | ···················· | ···················· | ···················· | ···················· |
| Q | 3401 hCoV-19/England/CAMC-AEAA9B/2020\|EPI_ISL_633120\|20 | ···················· | ···················· | ···················· | ···················· | ···················· | ···················· |
| Q | 3402 hCoV-19/England/CAMC-AEAFAB/2020\|EPI_ISL_633121\|20 | ···················· | ···················· | ···················· | ···················· | ···················· | ···················· |
| Q | 3403 hCoV-19/England/CAMC-AEAA7D/2020\|EPI_ISL_633122\|20 | ···················· | ···················· | ···················· | ···················· | ···················· | ···················· |
| Q | 3404 hCoV-19/England/CAMC-AEA9AD/2020\|EPI_ISL_633123\|20 | ···················· | ···················· | ···················· | ···················· | ······T············· | ···················· |
| Q | 3405 hCoV-19/England/CAMC-AEA819/2020\|EPI_ISL_633124\|20 | ···················· | ···················· | ···················· | ···················· | ···················· | ···················· |
| Q | 3406 hCoV-19/England/QEUH-AE540B/2020\|EPI_ISL_633125\|20 | ···················· | ···················· | ···················· | ···················· | ···················· | ···················· |
| Q | 3407 hCoV-19/England/CAMC-AEAECC/2020\|EPI_ISL_633126\|20 | ···················· | ···················· | ···················· | ···················· | ···················· | ···················· |
| Q | 3408 hCoV-19/England/CAMC-AEAF50/2020\|EPI_ISL_633127\|20 | ···················· | ···················· | ···················· | ···················· | ···················· | ···················· |
| Q | 3409 hCoV-19/England/CAMC-AEA6C4/2020\|EPI_ISL_633128\|20 | ···················· | ···················· | ···················· | ···················· | ···················· | ···················· |
| Q | 3410 hCoV-19/England/CAMC-AEB0C5/2020\|EPI_ISL_633129\|20 | ···················· | ···················· | ···················· | ···················· | ···················· | ···················· |
| Q | 3411 hCoV-19/England/CAMC-AEB098/2020\|EPI_ISL_633130\|20 | ···················· | ···················· | ···················· | ···················· | ···················· | ···················· |
| Q | 3412 hCoV-19/England/CAMC-AEAE80/2020\|EPI_ISL_633131\|20 | ···················· | ···················· | ···················· | ···················· | ···················· | ···················· |
| Q | 3413 hCoV-19/England/QEUH-AE4A28/2020\|EPI_ISL_633132\|20 | ···················· | ···················· | ···················· | ···················· | ···················· | ···················· |
| Q | 3414 hCoV-19/England/CAMC-AEB186/2020\|EPI_ISL_633135\|20 | ···················· | ···················· | ···················· | ···················· | ···················· | ···················· |
| Q | 3415 hCoV-19/England/CAMC-AEA9BC/2020\|EPI_ISL_633136\|20 | ···················· | ···················· | ···················· | ···················· | ···················· | ···················· |
| Q | 3416 hCoV-19/England/QEUH-AE526B/2020\|EPI_ISL_633137\|20 | ···················· | ···················· | ···················· | ···················· | ···················· | ···················· |
| Q | 3417 hCoV-19/England/QEUH-AE51D7/2020\|EPI_ISL_633138\|20 | ···················· | ···················· | ···················· | ···················· | ···················· | ···················· |
| Q | 3418 hCoV-19/England/QEUH-AE4E4A/2020\|EPI_ISL_633139\|20 | ···················· | ···················· | ···················· | ···················· | ···················· | ···················· |
| Q | 3419 hCoV-19/England/CAMC-AEA873/2020\|EPI_ISL_633140\|20 | ···················· | ···················· | ···················· | ···················· | ···················· | ···················· |
| Q | 3420 hCoV-19/England/CAMC-AEAB4D/2020\|EPI_ISL_633141\|20 | ···················· | ···················· | ···················· | ···················· | ···················· | ···················· |
| Q | 3421 hCoV-19/England/CAMC-AEB12C/2020\|EPI_ISL_633142\|20 | ···················· | ···················· | ···················· | ···················· | ···················· | ···················· |
| Q | 3422 hCoV-19/England/QEUH-AE4BAD/2020\|EPI_ISL_633143\|20 | ···················· | ···················· | ···················· | ···················· | ···················· | ···················· |
| Q | 3423 hCoV-19/England/QEUH-AE4958/2020\|EPI_ISL_633144\|20 | ···················· | ·A·················· | ···················· | ···················· | ···················· | ···················· |
| Q | 3424 hCoV-19/England/QEUH-AE52F2/2020\|EPI_ISL_633145\|20 | ···················· | ···················· | ···················· | ···················· | ···················· | ···················· |
| Q | 3425 hCoV-19/England/QEUH-AE53B3/2020\|EPI_ISL_633146\|20 | ···················· | ···················· | ···················· | ···················· | ···················· | ···················· |
| Q | 3426 hCoV-19/England/CAMC-AEAD74/2020\|EPI_ISL_633147\|20 | ···················· | ···················· | ···················· | ···················· | ···················· | ···················· |
| Q | 3427 hCoV-19/England/CAMC-AEAE26/2020\|EPI_ISL_633148\|20 | ···················· | ···················· | ···················· | ···················· | ···················· | ···················· |
| Q | 3428 hCoV-19/England/CAMC-AEAC86/2020\|EPI_ISL_633149\|20 | ···················· | ···················· | ···················· | ···················· | ···················· | ···················· |
| Q | 3429 hCoV-19/England/MILK-ACD409/2020\|EPI_ISL_634400\|20 | ···················· | ·A·················· | ···················· | ···················· | ···················· | ···A················ |
| Q | 3430 hCoV-19/England/MILK-ACC5CB/2020\|EPI_ISL_634401\|20 | ···················· | ···················· | ···················· | ···················· | ···················· | ···················· |
| Q | 3431 hCoV-19/England/MILK-ACC534/2020\|EPI_ISL_634402\|20 | ···················· | ···················· | ···················· | ···················· | ···················· | ···················· |
| Q | 3432 hCoV-19/England/MILK-AC7AEF/2020\|EPI_ISL_634403\|20 | ···················· | ···················· | ···················· | ···················· | ···················· | ···················· |
| Q | 3433 hCoV-19/England/MILK-ACC561/2020\|EPI_ISL_634404\|20 | ···················· | ···················· | ···················· | ···················· | ···················· | ···················· |
| Q | 3434 hCoV-19/England/MILK-ACD348/2020\|EPI_ISL_634405\|20 | ···················· | ···················· | ···················· | ···················· | ······T············· | ···················· |
| Q | 3435 hCoV-19/England/MILK-ACD384/2020\|EPI_ISL_634407\|20 | ···················· | ···················· | ···················· | ···················· | ···················· | ···················· |
| Q | 3436 hCoV-19/England/MILK-AC7C70/2020\|EPI_ISL_634409\|20 | ···················· | ···················· | ···················· | ···················· | ···················· | ···················· |
| Q | 3437 hCoV-19/England/MILK-AC7AB2/2020\|EPI_ISL_634410\|20 | ···················· | ···················· | ···················· | ···················· | ···················· | ···················· |
| Q | 3438 hCoV-19/England/MILK-AC796A/2020\|EPI_ISL_634411\|20 | ···················· | ···················· | ···················· | ···················· | ······T············· | ···················· |
| Q | 3439 hCoV-19/England/MILK-ACD6A9/2020\|EPI_ISL_634412\|20 | ···················· | ···················· | ···················· | ···················· | ···················· | ···················· |
| Q | 3440 hCoV-19/England/MILK-ACD72E/2020\|EPI_ISL_634413\|20 | ···················· | ···················· | ···················· | ···················· | ······T············· | ···················· |
| Q | 3441 hCoV-19/England/MILK-ACD4EB/2020\|EPI_ISL_634414\|20 | ···················· | ···················· | ···················· | ···················· | ···················· | ···················· |
| Q | 3442 hCoV-19/England/MILK-ACD1D5/2020\|EPI_ISL_634415\|20 | ···················· | ···················· | ···················· | ···················· | ···················· | ···················· |
| Q | 3443 hCoV-19/England/MILK-ACCEDF/2020\|EPI_ISL_634416\|20 | ···················· | ···················· | ···················· | ···················· | ···················· | ···················· |
| Q | 3444 hCoV-19/England/MILK-ACC877/2020\|EPI_ISL_634417\|20 | ···················· | ···················· | ···················· | ···················· | ···················· | ···················· |
| Q | 3445 hCoV-19/England/MILK-ACC5BC/2020\|EPI_ISL_634418\|20 | ···················· | ···················· | ···················· | ···················· | ···················· | ···················· |
| Q | 3446 hCoV-19/England/MILK-AC79F1/2020\|EPI_ISL_634419\|20 | ···················· | ···················· | ···················· | ···················· | ···················· | ···················· |
| Q | 3447 hCoV-19/England/MILK-ACCADB/2020\|EPI_ISL_634420\|20 | ···················· | ···················· | ···················· | ···················· | ···················· | ···················· |
| Q | 3448 hCoV-19/England/MILK-ACC68C/2020\|EPI_ISL_634421\|20 | ···················· | ···················· | ···················· | ···················· | ···················· | ···················· |
| Q | 3449 hCoV-19/England/MILK-ACD80D/2020\|EPI_ISL_634422\|20 | ···················· | ···················· | ···················· | ···················· | ···················· | ···················· |
| Q | 3450 hCoV-19/England/MILK-ACC437/2020\|EPI_ISL_634423\|20 | ···················· | ···················· | ···················· | ···················· | ···················· | ···················· |
| Q | 3451 hCoV-19/England/MILK-ACD5D9/2020\|EPI_ISL_634424\|20 | ···················· | ···················· | ···················· | ···················· | ···················· | ···················· |
| Q | 3452 hCoV-19/England/MILK-ACC4DD/2020\|EPI_ISL_634425\|20 | ···················· | ···················· | ···················· | ···················· | ···················· | ···················· |
| Q | 3453 hCoV-19/England/MILK-AC79E2/2020\|EPI_ISL_634426\|20 | ···················· | ···················· | ···················· | ···················· | ···················· | ···················· |
| Q | 3454 hCoV-19/England/MILK-ACD75B/2020\|EPI_ISL_634427\|20 | ···················· | ···················· | ···················· | ···················· | ···················· | ···················· |

| | | | | | | | |
|---|---|---|---|---|---|---|---|
| Q | 3455 hCoV-19/England/MILK-ACD24B/2020\|EPI_ISL_634429\|20 | ···················· | ···················· | ···················· | ···················· | ······T············· | ···················· |
| Q | 3456 hCoV-19/England/MILK-ACC4A0/2020\|EPI_ISL_634430\|20 | ···················· | ···················· | ···················· | ···················· | ···················· | ···················· |
| Q | 3457 hCoV-19/England/MILK-ACC83B/2020\|EPI_ISL_634431\|20 | ···················· | ···················· | ···················· | ···················· | ···················· | ···················· |
| Q | 3458 hCoV-19/England/MILK-ACC473/2020\|EPI_ISL_634432\|20 | ···················· | ···················· | ···················· | ···················· | ···················· | ···················· |
| Q | 3459 hCoV-19/England/MILK-AC7BEC/2020\|EPI_ISL_634433\|20 | ···················· | ···················· | ···················· | ···················· | ···················· | ···················· |
| Q | 3460 hCoV-19/England/MILK-ACCBC9/2020\|EPI_ISL_634434\|20 | ···················· | ···················· | ···················· | ···················· | ···················· | ···················· |
| Q | 3461 hCoV-19/England/MILK-ACD4DC/2020\|EPI_ISL_634435\|20 | ···················· | ···················· | ···················· | ···················· | ···················· | ···················· |
| Q | 3462 hCoV-19/England/MILK-ACCC20/2020\|EPI_ISL_634436\|20 | ···················· | ···················· | ···················· | ···················· | ···················· | ···················· |
| Q | 3463 hCoV-19/England/MILK-ACD9CE/2020\|EPI_ISL_634437\|20 | ···················· | ···················· | ···················· | ···················· | ···················· | ···················· |
| Q | 3464 hCoV-19/England/MILK-ACCC11/2020\|EPI_ISL_634438\|20 | ···················· | ···················· | ···················· | ···················· | ···················· | ···················· |
| Q | 3465 hCoV-19/England/MILK-AC787C/2020\|EPI_ISL_634439\|20 | ···················· | ···················· | ···················· | ···················· | ···················· | ···················· |
| Q | 3466 hCoV-19/England/MILK-ACCB9C/2020\|EPI_ISL_634440\|20 | ···················· | ···················· | ···················· | ···················· | ······T············· | ···················· |
| Q | 3467 hCoV-19/England/MILK-ACD3DF/2020\|EPI_ISL_634441\|20 | ···················· | ···················· | ···················· | ···················· | ···················· | ···················· |
| Q | 3468 hCoV-19/England/MILK-AC8116/2020\|EPI_ISL_634442\|20 | ···················· | ···················· | ···················· | ···················· | ···················· | ···················· |
| Q | 3469 hCoV-19/England/MILK-AC795B/2020\|EPI_ISL_634443\|20 | ···················· | ···················· | ···················· | ···················· | ······T············· | ···················· |
| Q | 3470 hCoV-19/England/MILK-AC7A76/2020\|EPI_ISL_634444\|20 | ···················· | ···················· | ···················· | ···················· | ···················· | ···················· |
| Q | 3471 hCoV-19/England/MILK-ACD4FA/2020\|EPI_ISL_634446\|20 | ···················· | ···················· | ···················· | ···················· | ···················· | ···················· |
| Q | 3472 hCoV-19/England/MILK-AC7988/2020\|EPI_ISL_634448\|20 | ···················· | ···················· | ···················· | ···················· | ···················· | ···················· |
| Q | 3473 hCoV-19/England/MILK-AC7B37/2020\|EPI_ISL_634449\|20 | ···················· | ···················· | ···················· | ···················· | ···················· | ···················· |
| Q | 3474 hCoV-19/England/MILK-ACD76A/2020\|EPI_ISL_634450\|20 | ···················· | ···················· | ···················· | ···················· | ···················· | ···················· |
| Q | 3475 hCoV-19/England/MILK-ACD630/2020\|EPI_ISL_634451\|20 | ···················· | ···················· | ···················· | ···················· | ···················· | ···················· |
| Q | 3476 hCoV-19/England/MILK-AC7A58/2020\|EPI_ISL_634452\|20 | ···················· | ···················· | ···················· | ···················· | ···················· | ···················· |
| Q | 3477 hCoV-19/England/MILK-ACD5BB/2020\|EPI_ISL_634453\|20 | ···················· | ··············C····· | ···················· | ···················· | ···················· | ···················· |
| Q | 3478 hCoV-19/England/MILK-AC75A2/2020\|EPI_ISL_634455\|20 | ···················· | ···················· | ···················· | ···················· | ······T············· | ···················· |
| Q | 3479 hCoV-19/England/MILK-AC7C25/2020\|EPI_ISL_634456\|20 | ···················· | ···················· | ···················· | ···················· | ···················· | ···················· |
| Q | 3480 hCoV-19/England/MILK-AC791F/2020\|EPI_ISL_634457\|20 | ···················· | ···················· | ···················· | ···················· | ······T············· | ···················· |
| Q | 3481 hCoV-19/England/MILK-ACD58E/2020\|EPI_ISL_634458\|20 | ···················· | ···················· | ···················· | ···················· | ······T············· | ···················· |
| Q | 3482 hCoV-19/England/MILK-ACC613/2020\|EPI_ISL_634459\|20 | ···················· | ···················· | ···················· | ···················· | ···················· | ···················· |
| Q | 3483 hCoV-19/England/MILK-ACC82C/2020\|EPI_ISL_634461\|20 | ···················· | ···················· | ···················· | ···················· | ···················· | ···················· |
| Q | 3484 hCoV-19/England/MILK-ACD6D6/2020\|EPI_ISL_634463\|20 | ···················· | ···················· | ···················· | ···················· | ······T············· | ···················· |
| Q | 3485 hCoV-19/England/MILK-AC78B8/2020\|EPI_ISL_634464\|20 | ···················· | ···················· | ···················· | ···················· | ······T············· | ···················· |
| Q | 3486 hCoV-19/England/MILK-ACD82B/2020\|EPI_ISL_634466\|20 | ···················· | ···················· | ···················· | ···················· | ···················· | ···················· |
| Q | 3487 hCoV-19/England/MILK-ACC7C5/2020\|EPI_ISL_634467\|20 | ···················· | ···················· | ···················· | ···················· | ···················· | ···················· |
| Q | 3488 hCoV-19/England/MILK-AC7979/2020\|EPI_ISL_634468\|20 | ···················· | ···················· | ···················· | ···················· | ···················· | ···················· |
| Q | 3489 hCoV-19/England/MILK-AC7900/2020\|EPI_ISL_634469\|20 | ···················· | ···················· | ···················· | ···················· | ···················· | ···················· |
| Q | 3490 hCoV-19/England/MILK-ACD3A2/2020\|EPI_ISL_634470\|20 | ···················· | ···················· | ···················· | ···················· | ······T············· | ···················· |
| Q | 3491 hCoV-19/England/MILK-AC8152/2020\|EPI_ISL_634471\|20 | ···················· | ···················· | ···················· | ···················· | ···················· | ···················· |
| Q | 3492 hCoV-19/England/MILK-AC7BDD/2020\|EPI_ISL_634472\|20 | ···················· | ···················· | ···················· | ···················· | ···················· | ···················· |
| Q | 3493 hCoV-19/England/MILK-ACD1A8/2020\|EPI_ISL_634473\|20 | ···················· | ···················· | ···················· | ···················· | ···················· | ···················· |
| Q | 3494 hCoV-19/England/MILK-ACCAEA/2020\|EPI_ISL_634474\|20 | ···················· | ···················· | ···················· | ···················· | ···················· | ···················· |
| Q | 3495 hCoV-19/England/MILK-AC7B28/2020\|EPI_ISL_634476\|20 | ···················· | ·A·················· | ···················· | ···················· | ···················· | ···A················ |
| Q | 3496 hCoV-19/England/MILK-ACCF09/2020\|EPI_ISL_634477\|20 | ···················· | ·A·················· | ···················· | ···················· | ···················· | ···A················ |
| Q | 3497 hCoV-19/England/MILK-ACD7F1/2020\|EPI_ISL_634478\|20 | ···················· | ···················· | ···················· | ···················· | ···················· | ···················· |
| Q | 3498 hCoV-19/England/MILK-ACD375/2020\|EPI_ISL_634479\|20 | ···················· | ···················· | ···················· | ···················· | ···················· | ···················· |
| Q | 3499 hCoV-19/USA/CA-ALSR-2829/2020\|EPI_ISL_635701\|2020- | ···················· | ·A·················· | ···················· | ···················· | ···················· | ···················· |
| Q | 3500 hCoV-19/USA/CA-ALSR-2831/2020\|EPI_ISL_635702\|2020- | ···················· | ·A·················· | ···················· | ···················· | ···················· | ···················· |
| Q | 3501 hCoV-19/USA/CA-ALSR-2832/2020\|EPI_ISL_635703\|2020- | ···················· | ···················· | ···················· | ···················· | ···················· | ···················· |
| Q | 3502 hCoV-19/USA/CA-ALSR-2834/2020\|EPI_ISL_635704\|2020- | ···················· | ···················· | ···················· | ···················· | ···················· | ···················· |
| Q | 3503 hCoV-19/USA/CA-ALSR-2838/2020\|EPI_ISL_635705\|2020- | ···················· | ···················· | ···················· | ···················· | ···················· | ···················· |
| Q | 3504 hCoV-19/USA/CA-ALSR-2840/2020\|EPI_ISL_635706\|2020- | ···················· | ···················· | ···················· | ···················· | ···················· | ···················· |
| Q | 3505 hCoV-19/USA/CA-ALSR-2842/2020\|EPI_ISL_635708\|2020- | ···················· | ···················· | ···················· | ···················· | ···················· | ···················· |
| Q | 3506 hCoV-19/USA/CA-ALSR-2843/2020\|EPI_ISL_635709\|2020- | ···················· | ·A·················· | ···················· | ···················· | ···················· | ···················· |
| Q | 3507 hCoV-19/USA/CA-ALSR-2844/2020\|EPI_ISL_635710\|2020- | ···················· | ···················· | ···················· | ···················· | ···················· | ···················· |
| Q | 3508 hCoV-19/USA/CA-ALSR-2849/2020\|EPI_ISL_635711\|2020- | ···················· | ·A·················· | ···················· | ···················· | ···················· | ···················· |
| Q | 3509 hCoV-19/USA/CA-ALSR-2850/2020\|EPI_ISL_635712\|2020- | ···················· | ·A·················· | ···················· | ···················· | ···················· | ···················· |
| Q | 3510 hCoV-19/USA/CA-ALSR-2854/2020\|EPI_ISL_635713\|2020- | ···················· | ···················· | ···················· | ···················· | ···················· | ···················· |
| Q | 3511 hCoV-19/USA/CA-ALSR-2855/2020\|EPI_ISL_635714\|2020- | ···················· | ·A·················· | ···················· | ···················· | ···················· | ···················· |
| Q | 3512 hCoV-19/USA/CA-ALSR-2857/2020\|EPI_ISL_635716\|2020- | ···················· | ·A·················· | ···················· | ···················· | ···················· | ···················· |
| Q | 3513 hCoV-19/USA/CA-ALSR-2872/2020\|EPI_ISL_635719\|2020- | ···················· | ·A·················· | ···················· | ···················· | ···················· | ···················· |
| Q | 3514 hCoV-19/USA/CA-ALSR-2874/2020\|EPI_ISL_635720\|2020- | ···················· | ·A·················· | ···················· | ···················· | ···················· | ···················· |
| Q | 3515 hCoV-19/USA/CA-ALSR-2875/2020\|EPI_ISL_635721\|2020- | ···················· | ·A·················· | ···················· | ···················· | ···················· | ···················· |
| Q | 3516 hCoV-19/USA/CA-ALSR-2880/2020\|EPI_ISL_635722\|2020- | ···················· | ··C················· | ···················· | ···················· | ···················· | ···················· |
| Q | 3517 hCoV-19/USA/CA-ALSR-2890/2020\|EPI_ISL_635724\|2020- | ···················· | ·A·················· | ···················· | ···················· | ···················· | ···················· |
| Q | 3518 hCoV-19/USA/CA-ALSR-2892/2020\|EPI_ISL_635725\|2020- | ···················· | ·A·················· | ···················· | ···················· | ···················· | ···················· |
| Q | 3519 hCoV-19/USA/CA-ALSR-2896/2020\|EPI_ISL_635726\|2020- | ···················· | ···················· | ···················· | ···················· | ···················· | ···················· |
| Q | 3520 hCoV-19/USA/CA-ALSR-2901/2020\|EPI_ISL_635727\|2020- | ···················· | ·A·················· | ···················· | ···················· | ···················· | ···················· |
| Q | 3521 hCoV-19/USA/CA-ALSR-2904/2020\|EPI_ISL_635729\|2020- | ···················· | ···················· | ···················· | ···················· | ···················· | ···················· |
| Q | 3522 hCoV-19/USA/CA-ALSR-2905/2020\|EPI_ISL_635730\|2020- | ···················· | ·A·················· | ···················· | ···················· | ···················· | ···················· |
| Q | 3523 hCoV-19/USA/CA-ALSR-2909/2020\|EPI_ISL_635731\|2020- | ···················· | ·A·················· | ···················· | ···················· | ···················· | ···················· |
| Q | 3524 hCoV-19/USA/CA-ALSR-2915/2020\|EPI_ISL_635732\|2020- | ···················· | ·A·················· | ···················· | ···················· | ···················· | ···················· |
| Q | 3525 hCoV-19/USA/CA-ALSR-2922/2020\|EPI_ISL_635733\|2020- | ···················· | ···················· | ···················· | ···················· | ···················· | ···················· |
| Q | 3526 hCoV-19/USA/CA-ALSR-2927/2020\|EPI_ISL_635734\|2020- | ···················· | ···················· | ···················· | ···················· | ···················· | ···················· |
| Q | 3527 hCoV-19/USA/CA-ALSR-2929/2020\|EPI_ISL_635736\|2020- | ···················· | ·A·················· | ···················· | ···················· | ···················· | ···················· |
| Q | 3528 hCoV-19/USA/CA-ALSR-2932/2020\|EPI_ISL_635737\|2020- | ···················· | ·A·················· | ···················· | ···················· | ···················· | ···················· |
| Q | 3529 hCoV-19/USA/CA-ALSR-2936/2020\|EPI_ISL_635739\|2020- | ···················· | ·A·················· | ···················· | ···················· | ···················· | ···················· |
| Q | 3530 hCoV-19/USA/CA-ALSR-2937/2020\|EPI_ISL_635740\|2020- | ···················· | ·A·················· | ···················· | ···················· | ···················· | ···················· |
| Q | 3531 hCoV-19/USA/CA-ALSR-2938/2020\|EPI_ISL_635741\|2020- | ···················· | ·A·················· | ···················· | ···················· | ···················· | ···················· |
| Q | 3532 hCoV-19/USA/CA-ALSR-2939/2020\|EPI_ISL_635742\|2020- | ···················· | ···················· | ···················· | ···················· | ···················· | ···················· |
| Q | 3533 hCoV-19/USA/CA-ALSR-2941/2020\|EPI_ISL_635743\|2020- | ···················· | ···················· | ···················· | ···················· | ···················· | ···················· |
| Q | 3534 hCoV-19/USA/CA-ALSR-2943/2020\|EPI_ISL_635744\|2020- | ···················· | ·A·················· | ···················· | ···················· | ···················· | ···················· |

| | | | | | | | |
|---|---|---|---|---|---|---|---|
| Q | 3535 hCoV-19/USA/CA-ALSR-2947/2020\|EPI_ISL_635745\|2020- | ···················· | ·A·················· | ···················· | ···················· | ···················· | ···················· |
| Q | 3536 hCoV-19/USA/CA-ALSR-2949/2020\|EPI_ISL_635746\|2020- | ···················· | ·A·················· | ···················· | ···················· | ···················· | ···················· |
| Q | 3537 hCoV-19/USA/CA-ALSR-2955/2020\|EPI_ISL_635747\|2020- | ···················· | ·A·················· | ···················· | ···················· | ···················· | ···················· |
| Q | 3538 hCoV-19/USA/CA-ALSR-2958/2020\|EPI_ISL_635748\|2020- | ···················· | ···················· | ···················· | ···················· | ···················· | ···················· |
| Q | 3539 hCoV-19/USA/CA-ALSR-2959/2020\|EPI_ISL_635749\|2020- | ···················· | ·A·················· | ···················· | ···················· | ···················· | ···················· |
| Q | 3540 hCoV-19/USA/CA-ALSR-2960/2020\|EPI_ISL_635750\|2020- | ···················· | ·A·················· | ···················· | ···················· | ···················· | ···················· |
| Q | 3541 hCoV-19/USA/CA-ALSR-2961/2020\|EPI_ISL_635751\|2020- | ···················· | ·A·················· | ···················· | ···················· | ···················· | ···················· |
| Q | 3542 hCoV-19/USA/CA-ALSR-2962/2020\|EPI_ISL_635752\|2020- | ···················· | ·A·················· | ···················· | ···················· | ···················· | ···················· |
| Q | 3543 hCoV-19/USA/CA-ALSR-2963/2020\|EPI_ISL_635753\|2020- | ···················· | ·A·················· | ···················· | ···················· | ···················· | ···················· |
| Q | 3544 hCoV-19/USA/CA-ALSR-2964/2020\|EPI_ISL_635754\|2020- | ···················· | ·A·················· | ···················· | ···················· | ···················· | ···················· |
| Q | 3545 hCoV-19/USA/CA-ALSR-2965/2020\|EPI_ISL_635755\|2020- | ···················· | ·A·················· | ···················· | ···················· | ···················· | ···················· |
| Q | 3546 hCoV-19/USA/CA-ALSR-2966/2020\|EPI_ISL_635756\|2020- | ···················· | ·A·················· | ···················· | ···················· | ···················· | ···················· |
| Q | 3547 hCoV-19/USA/CA-ALSR-2967/2020\|EPI_ISL_635757\|2020- | ···················· | ·A·················· | ···················· | ···················· | ···················· | ···················· |
| Q | 3548 hCoV-19/USA/CA-ALSR-2968/2020\|EPI_ISL_635758\|2020- | ···················· | ·A·················· | ···················· | ···················· | ···················· | ···················· |
| Q | 3549 hCoV-19/USA/CA-ALSR-2969/2020\|EPI_ISL_635759\|2020- | ···················· | ·A·················· | ···················· | ···················· | ···················· | ···················· |
| Q | 3550 hCoV-19/USA/CA-ALSR-2970/2020\|EPI_ISL_635760\|2020- | ···················· | ·A·················· | ···················· | ···················· | ···················· | ···················· |
| Q | 3551 hCoV-19/USA/CA-ALSR-2971/2020\|EPI_ISL_635761\|2020- | ···················· | ·A·················· | ···················· | ···················· | ···················· | ···················· |
| Q | 3552 hCoV-19/USA/CA-ALSR-2972/2020\|EPI_ISL_635762\|2020- | ···················· | ·A·················· | ···················· | ···················· | ···················· | ···················· |
| Q | 3553 hCoV-19/USA/CA-ALSR-2973/2020\|EPI_ISL_635763\|2020- | ···················· | ·A·················· | ···················· | ···················· | ···················· | ···················· |
| Q | 3554 hCoV-19/USA/CA-ALSR-2974/2020\|EPI_ISL_635764\|2020- | ···················· | ·A·················· | ···················· | ···················· | ···················· | ···················· |
| Q | 3555 hCoV-19/USA/CA-ALSR-2975/2020\|EPI_ISL_635765\|2020- | ···················· | ·A·················· | ···················· | ···················· | ···················· | ···················· |
| Q | 3556 hCoV-19/USA/CA-ALSR-2976/2020\|EPI_ISL_635766\|2020- | ···················· | ·A·················· | ···················· | ···················· | ···················· | ···················· |
| Q | 3557 hCoV-19/USA/CA-ALSR-2977/2020\|EPI_ISL_635767\|2020- | ···················· | ·A·················· | ···················· | ···················· | ···················· | ···················· |
| Q | 3558 hCoV-19/USA/CA-ALSR-2979/2020\|EPI_ISL_635768\|2020- | ···················· | ·A·················· | ···················· | ···················· | ···················· | ···················· |
| Q | 3559 hCoV-19/USA/CA-ALSR-2980/2020\|EPI_ISL_635769\|2020- | ···················· | ·A·················· | ···················· | ···················· | ···················· | ···················· |
| Q | 3560 hCoV-19/USA/CA-ALSR-2981/2020\|EPI_ISL_635770\|2020- | ···················· | ·A·················· | ···················· | ···················· | ···················· | ···················· |
| Q | 3561 hCoV-19/USA/CA-ALSR-2982/2020\|EPI_ISL_635771\|2020- | ···················· | ·A·················· | ···················· | ···················· | ···················· | ···················· |
| Q | 3562 hCoV-19/USA/CA-ALSR-2984/2020\|EPI_ISL_635772\|2020- | ···················· | ·A·················· | ···················· | ···················· | ···················· | ···················· |
| Q | 3563 hCoV-19/USA/CA-ALSR-2985/2020\|EPI_ISL_635773\|2020- | ···················· | ·A·················· | ···················· | ···················· | ···················· | ···················· |
| Q | 3564 hCoV-19/USA/CA-ALSR-2987/2020\|EPI_ISL_635774\|2020- | ···················· | ·A·················· | ···················· | ···················· | ···················· | ···················· |
| Q | 3565 hCoV-19/USA/CA-ALSR-2988/2020\|EPI_ISL_635775\|2020- | ···················· | ·A·················· | ···················· | ···················· | ···················· | ···················· |
| Q | 3566 hCoV-19/USA/CA-ALSR-2989/2020\|EPI_ISL_635776\|2020- | ···················· | ·A·················· | ···················· | ···················· | ···················· | ···················· |
| Q | 3567 hCoV-19/Jordan/IR-ALSR-2578/2020\|EPI_ISL_635777\|20 | ···················· | ···················· | ···················· | ···················· | ···················· | ···················· |
| Q | 3568 hCoV-19/Jordan/AM-ALSR-2579/2020\|EPI_ISL_635778\|20 | ···················· | ···················· | ···················· | ···················· | ···················· | ···················· |
| Q | 3569 hCoV-19/Jordan/MA-ALSR-2581/2020\|EPI_ISL_635779\|20 | ···················· | ·A·················· | ···················· | ···················· | ···················· | ···················· |
| Q | 3570 hCoV-19/Jordan/MA-ALSR-2583/2020\|EPI_ISL_635780\|20 | ···················· | ·A·················· | ···················· | ···················· | ···················· | ···················· |
| Q | 3571 hCoV-19/Jordan/AM-ALSR-2585/2020\|EPI_ISL_635781\|20 | ···················· | ···················· | ···················· | ···················· | ···················· | ···················· |
| Q | 3572 hCoV-19/Jordan/AM-ALSR-2586/2020\|EPI_ISL_635782\|20 | ···················· | ···················· | ···················· | ···················· | ···················· | ···················· |
| Q | 3573 hCoV-19/USA/CA-ALSR-3012/2020\|EPI_ISL_635786\|2020- | ···················· | ·A·················· | ···················· | ···················· | ···················· | ···················· |
| Q | 3574 hCoV-19/USA/CA-ALSR-3013/2020\|EPI_ISL_635787\|2020- | ···················· | ·A·················· | ···················· | ···················· | ···················· | ···················· |
| Q | 3575 hCoV-19/USA/CA-ALSR-3015/2020\|EPI_ISL_635788\|2020- | ···················· | ·A·················· | ···················· | ···················· | ···················· | ···················· |
| Q | 3576 hCoV-19/USA/CA-ALSR-3018/2020\|EPI_ISL_635790\|2020- | ···················· | ·A·················· | ···················· | ···················· | ···················· | ···················· |
| Q | 3577 hCoV-19/USA/CA-ALSR-3019/2020\|EPI_ISL_635791\|2020- | ···················· | ···················· | ···················· | ···················· | ···················· | ···················· |
| Q | 3578 hCoV-19/USA/CA-ALSR-3021/2020\|EPI_ISL_635792\|2020- | ···················· | ·A·················· | ···················· | ···················· | ···················· | ···················· |
| Q | 3579 hCoV-19/USA/CA-ALSR-3022/2020\|EPI_ISL_635793\|2020- | ···················· | ·A·················· | ···················· | ···················· | ···················· | ···················· |
| Q | 3580 hCoV-19/USA/CA-ALSR-3023/2020\|EPI_ISL_635794\|2020- | ···················· | ·A·················· | ···················· | ···················· | ···················· | ···················· |
| Q | 3581 hCoV-19/USA/CA-ALSR-3024/2020\|EPI_ISL_635795\|2020- | ···················· | ···················· | ···················· | ···················· | ···················· | ···················· |
| Q | 3582 hCoV-19/USA/CA-ALSR-3026/2020\|EPI_ISL_635796\|2020- | ···················· | ·A·················· | ···················· | ···················· | ···················· | ···················· |
| Q | 3583 hCoV-19/USA/CA-ALSR-3027/2020\|EPI_ISL_635797\|2020- | ···················· | ·A·················· | ···················· | ···················· | ···················· | ···················· |
| Q | 3584 hCoV-19/USA/CA-ALSR-3029/2020\|EPI_ISL_635799\|2020- | ···················· | ·A·················· | ···················· | ···················· | ···················· | ···················· |
| Q | 3585 hCoV-19/England/MILK-A18E01/2020\|EPI_ISL_627203\|20 | ···················· | ···················· | ···················· | ···················· | ···················· | ···················· |
| Q | 3587 hCoV-19/Wales/PHWC-485913/2020\|EPI_ISL_627205\|2020 | ···················· | ···················· | ···················· | ···················· | ···················· | ···················· |
| Q | 3588 hCoV-19/Wales/PHWC-485F3F/2020\|EPI_ISL_627206\|2020 | ···················· | ···················· | ···················· | ···················· | ···················· | ···················· |
| Q | 3589 hCoV-19/Wales/PHWC-485E14/2020\|EPI_ISL_627207\|2020 | ···················· | ···················· | ···················· | ···················· | ···················· | ···················· |
| Q | 3590 hCoV-19/Scotland/CVR5135/2020\|EPI_ISL_627209\|2020- | ···················· | ···················· | ···················· | ···················· | ···················· | ···················· |
| Q | 3591 hCoV-19/Wales/PHWC-48470B/2020\|EPI_ISL_627230\|2020 | ···················· | ···················· | ···················· | ···················· | ···················· | ···················· |
| Q | 3592 hCoV-19/Wales/PHWC-4849E7/2020\|EPI_ISL_627231\|2020 | ···················· | ···················· | ···················· | ···················· | ···················· | ···················· |
| Q | 3593 hCoV-19/Wales/PHWC-484A02/2020\|EPI_ISL_627232\|2020 | ···················· | ···················· | ···················· | ···················· | ···················· | ···················· |
| Q | 3594 hCoV-19/Wales/PHWC-484A5D/2020\|EPI_ISL_627233\|2020 | ···················· | ···················· | ···················· | ···················· | ···················· | ···················· |
| Q | 3595 hCoV-19/Wales/PHWC-484BD2/2020\|EPI_ISL_627234\|2020 | ···················· | ···················· | ···················· | ···················· | ···················· | ···················· |
| Q | 3596 hCoV-19/Wales/PHWC-484E33/2020\|EPI_ISL_627235\|2020 | ···················· | ···················· | ···················· | ···················· | ···················· | ···················· |
| Q | 3597 hCoV-19/Wales/PHWC-484FE5/2020\|EPI_ISL_627236\|2020 | ···················· | ···················· | ···················· | ···················· | ···················· | ···················· |
| Q | 3598 hCoV-19/Wales/PHWC-4852EB/2020\|EPI_ISL_627237\|2020 | ···················· | ···················· | ···················· | ···················· | ···················· | ···················· |
| Q | 3599 hCoV-19/Wales/PHWC-4853D9/2020\|EPI_ISL_627238\|2020 | ···················· | ···················· | ···················· | ···················· | ···················· | ···················· |
| Q | 3600 hCoV-19/Wales/PHWC-48548B/2020\|EPI_ISL_627239\|2020 | ···················· | ···················· | ···················· | ···················· | ···················· | ···················· |
| Q | 3601 hCoV-19/Wales/PHWC-485685/2020\|EPI_ISL_627240\|2020 | ···················· | ···················· | ···················· | ···················· | ···················· | ···················· |
| Q | 3602 hCoV-19/Wales/PHWC-485834/2020\|EPI_ISL_627241\|2020 | ···················· | ···················· | ···················· | ···················· | ···················· | ···················· |
| Q | 3604 hCoV-19/Wales/PHWC-48597D/2020\|EPI_ISL_627243\|2020 | ···················· | ···················· | ···················· | ···················· | ···················· | ···················· |
| Q | 3605 hCoV-19/Wales/PHWC-485C92/2020\|EPI_ISL_627244\|2020 | ···················· | ···················· | ···················· | ···················· | ···················· | ···················· |
| Q | 3606 hCoV-19/Wales/PHWC-485D71/2020\|EPI_ISL_627245\|2020 | ···················· | ···················· | ···················· | ···················· | ···················· | ···················· |
| Q | 3607 hCoV-19/Wales/PHWC-485D9F/2020\|EPI_ISL_627246\|2020 | ···················· | ···················· | ···················· | ···················· | ···················· | ···················· |
| Q | 3608 hCoV-19/Wales/PHWC-485FA8/2020\|EPI_ISL_627247\|2020 | ···················· | ···················· | ···················· | ···················· | ···················· | ···················· |
| Q | 3609 hCoV-19/Wales/PHWC-486068/2020\|EPI_ISL_627248\|2020 | ···················· | ···················· | ···················· | ···················· | ···················· | ···················· |
| Q | 3610 hCoV-19/Wales/PHWC-486138/2020\|EPI_ISL_627249\|2020 | ···················· | ···················· | ···················· | ···················· | ···················· | ···················· |
| Q | 3611 hCoV-19/Scotland/QEUH-A32250/2020\|EPI_ISL_627250\|2 | ···················· | ···················· | ···················· | ···················· | ···················· | ···················· |
| Q | 3612 hCoV-19/England/QEUH-A328A9/2020\|EPI_ISL_627251\|20 | ···················· | ···················· | ···················· | ···················· | ···················· | ···················· |
| Q | 3613 hCoV-19/Scotland/QEUH-A33565/2020\|EPI_ISL_627252\|2 | ···················· | ···················· | ···················· | ···················· | ···················· | ···················· |
| Q | 3614 hCoV-19/Wales/QEUH-A335A1/2020\|EPI_ISL_627253\|2020 | ···················· | ···················· | ···················· | ···················· | ···················· | ···················· |
| Q | 3615 hCoV-19/Scotland/QEUH-A3369F/2020\|EPI_ISL_627254\|2 | ···················· | ···················· | ···················· | ···················· | ···················· | ···················· |
| Q | 3616 hCoV-19/England/QEUH-A49A6D/2020\|EPI_ISL_627255\|20 | ···················· | ···················· | ···················· | ···················· | ···················· | ···················· |

| | | | | | | | |
|---|---|---|---|---|---|---|---|
| Q | 3617 hCoV-19/England/QEUH-A4AF12/2020\|EPI_ISL_627256\|20 | ···················· | ···················· | ···················· | ···················· | ···················· | ···················· |
| Q | 3618 hCoV-19/Scotland/QEUH-A4B97D/2020\|EPI_ISL_627257\|2 | ···················· | ···················· | ···················· | ···················· | ···················· | ···················· |
| Q | 3619 hCoV-19/England/QEUH-A4C095/2020\|EPI_ISL_627258\|20 | ···················· | ···················· | ···················· | ···················· | ···················· | ···················· |
| Q | 3620 hCoV-19/Scotland/QEUH-A4C1CF/2020\|EPI_ISL_627259\|2 | ···················· | ···················· | ···················· | ···················· | ···················· | ···················· |
| Q | 3621 hCoV-19/England/QEUH-A61D57/2020\|EPI_ISL_627260\|20 | ···················· | ···················· | ···················· | ···················· | ···················· | ···················· |
| Q | 3622 hCoV-19/England/QEUH-A63BF1/2020\|EPI_ISL_627261\|20 | ···················· | ···················· | ···················· | ···················· | ···················· | ···················· |
| Q | 3623 hCoV-19/England/QEUH-A63F04/2020\|EPI_ISL_627262\|20 | ···················· | ···················· | ···················· | ···················· | ···················· | ···················· |
| Q | 3624 hCoV-19/Scotland/QEUH-A4B9E6/2020\|EPI_ISL_628500\|2 | ···················· | ···················· | ···················· | ···················· | ···················· | ···················· |
| Q | 3625 hCoV-19/Scotland/QEUH-A4B9F5/2020\|EPI_ISL_628501\|2 | ···················· | ···················· | ···················· | ···················· | ···················· | ···················· |
| Q | 3626 hCoV-19/Scotland/QEUH-A4BA01/2020\|EPI_ISL_628502\|2 | ···················· | ···················· | ···················· | ···················· | ···················· | ···················· |
| Q | 3627 hCoV-19/Scotland/QEUH-A4BA10/2020\|EPI_ISL_628503\|2 | ···················· | ···················· | ···················· | ···················· | ···················· | ···················· |
| Q | 3628 hCoV-19/Scotland/QEUH-A4BA2F/2020\|EPI_ISL_628504\|2 | ···················· | ···················· | ···················· | ···················· | ···················· | ···················· |
| Q | 3629 hCoV-19/Scotland/QEUH-A4BA5C/2020\|EPI_ISL_628505\|2 | ···················· | ·A·················· | ···················· | ···················· | ···················· | ···················· |
| Q | 3630 hCoV-19/Scotland/QEUH-A4BA6B/2020\|EPI_ISL_628506\|2 | ···················· | ···················· | ···················· | ···················· | ···················· | ···················· |
| Q | 3631 hCoV-19/Scotland/QEUH-A4BA7A/2020\|EPI_ISL_628507\|2 | ···················· | ···················· | ···················· | ···················· | ···················· | ···················· |
| Q | 3632 hCoV-19/Scotland/QEUH-A4BAE3/2020\|EPI_ISL_628508\|2 | ···················· | ···················· | ···················· | ···················· | ···················· | ···················· |
| Q | 3633 hCoV-19/England/QEUH-A4BB2C/2020\|EPI_ISL_628510\|20 | ···················· | ···················· | ···················· | ···················· | ···················· | ···················· |
| Q | 3634 hCoV-19/Scotland/QEUH-A4BB95/2020\|EPI_ISL_628511\|2 | ···················· | ···················· | ···················· | ···················· | ···················· | ···················· |
| Q | 3635 hCoV-19/Scotland/QEUH-A4BBB3/2020\|EPI_ISL_628512\|2 | ···················· | ···················· | ···················· | ···················· | ···················· | ···················· |
| Q | 3636 hCoV-19/Scotland/QEUH-A4BBC2/2020\|EPI_ISL_628513\|2 | ···················· | ···················· | ···················· | ···················· | ···················· | ···················· |
| Q | 3637 hCoV-19/Scotland/QEUH-A4BBE0/2020\|EPI_ISL_628515\|2 | ···················· | ···················· | ···················· | ···················· | ···················· | ···················· |
| Q | 3638 hCoV-19/Scotland/QEUH-A4BBFF/2020\|EPI_ISL_628516\|2 | ···················· | ···················· | ···················· | ···················· | ···················· | ···················· |
| Q | 3639 hCoV-19/Scotland/QEUH-A4BC0B/2020\|EPI_ISL_628517\|2 | ···················· | ···················· | ···················· | ···················· | ···················· | ···················· |
| Q | 3640 hCoV-19/Scotland/QEUH-A4BC29/2020\|EPI_ISL_628518\|2 | ···················· | ···················· | ···················· | ···················· | ···················· | ···················· |
| Q | 3641 hCoV-19/England/QEUH-A4BC47/2020\|EPI_ISL_628519\|20 | ···················· | ···················· | ···················· | ···················· | ···················· | ···················· |
| Q | 3642 hCoV-19/England/QEUH-A4BF7B/2020\|EPI_ISL_628534\|20 | ···················· | ···················· | ···················· | ···················· | ···················· | ···················· |
| Q | 3643 hCoV-19/England/QEUH-A4BFC6/2020\|EPI_ISL_628535\|20 | ···················· | ···················· | ···················· | ···················· | ···················· | ···················· |
| Q | 3644 hCoV-19/Wales/QEUH-A4BFD5/2020\|EPI_ISL_628536\|2020 | ···················· | ···················· | ···················· | ···················· | ···················· | ···················· |
| Q | 3645 hCoV-19/England/QEUH-A4BFF3/2020\|EPI_ISL_628537\|20 | ···················· | ···················· | ···················· | ···················· | ···················· | ···················· |
| Q | 3646 hCoV-19/England/QEUH-A4C03B/2020\|EPI_ISL_628538\|20 | ···················· | ···················· | ···················· | ···················· | ···················· | ···················· |
| Q | 3647 hCoV-19/England/QEUH-A4C04A/2020\|EPI_ISL_628539\|20 | ···················· | ···················· | ···················· | ···················· | ···················· | ···················· |
| Q | 3648 hCoV-19/England/QEUH-A4C2F9/2020\|EPI_ISL_628560\|20 | ···················· | ···················· | ···················· | ···················· | ···················· | ···················· |
| Q | 3649 hCoV-19/Scotland/QEUH-A4C305/2020\|EPI_ISL_628561\|2 | ···················· | ···················· | ···················· | ···················· | ···················· | ···················· |
| Q | 3650 hCoV-19/Scotland/QEUH-A4C341/2020\|EPI_ISL_628562\|2 | ···················· | ···················· | ···················· | ···················· | ···················· | ···················· |
| Q | 3651 hCoV-19/Scotland/QEUH-A4C350/2020\|EPI_ISL_628563\|2 | ···················· | ···················· | ···················· | ···················· | ···················· | ···················· |
| Q | 3652 hCoV-19/Scotland/QEUH-A4C36F/2020\|EPI_ISL_628564\|2 | ···················· | ···················· | ···················· | ···················· | ···················· | ···················· |
| Q | 3653 hCoV-19/Scotland/QEUH-A4C3BA/2020\|EPI_ISL_628565\|2 | ···················· | ···················· | ···················· | ···················· | ···················· | ···················· |
| Q | 3654 hCoV-19/Scotland/QEUH-A4C3C9/2020\|EPI_ISL_628566\|2 | ···················· | ···················· | ···················· | ···················· | ···················· | ···················· |
| Q | 3655 hCoV-19/England/QEUH-A4C3D8/2020\|EPI_ISL_628567\|20 | ···················· | ···················· | ···················· | ···················· | ···················· | ···················· |
| Q | 3656 hCoV-19/England/QEUH-A4C420/2020\|EPI_ISL_628568\|20 | ···················· | ···················· | ···················· | ···················· | ······T············· | ···············A···· |
| Q | 3657 hCoV-19/Scotland/QEUH-A4C4B7/2020\|EPI_ISL_628569\|2 | ···················· | ···················· | ···················· | ···················· | ···················· | ···················· |
| Q | 3658 hCoV-19/Wales/QEUH-A4C4C6/2020\|EPI_ISL_628570\|2020 | ···················· | ···················· | ···················· | ···················· | ···················· | ···················· |
| Q | 3659 hCoV-19/England/QEUH-A4C61B/2020\|EPI_ISL_628571\|20 | ···················· | ···················· | ···················· | ···················· | ···················· | ···················· |
| Q | 3660 hCoV-19/England/QEUH-A4C772/2020\|EPI_ISL_628572\|20 | ···················· | ···················· | ···················· | ···················· | ···················· | ···················· |
| Q | 3661 hCoV-19/England/QEUH-A4C781/2020\|EPI_ISL_628573\|20 | ···················· | ···················· | ···················· | ···················· | ···················· | ···················· |
| Q | 3662 hCoV-19/England/QEUH-A4C7CD/2020\|EPI_ISL_628574\|20 | ···················· | ···················· | ···················· | ···················· | ···················· | ···················· |
| Q | 3663 hCoV-19/England/QEUH-A4C8E8/2020\|EPI_ISL_628575\|20 | ···················· | ···················· | ···················· | ···················· | ···················· | ···················· |
| Q | 3664 hCoV-19/England/QEUH-A4C8F7/2020\|EPI_ISL_628576\|20 | ···················· | ·A·················· | ···················· | ···················· | ···················· | ···A················ |
| Q | 3665 hCoV-19/England/QEUH-A4C912/2020\|EPI_ISL_628577\|20 | ···················· | ···················· | ···················· | ···················· | ···················· | ···················· |
| Q | 3666 hCoV-19/England/QEUH-A4C921/2020\|EPI_ISL_628578\|20 | ···················· | ···················· | ···················· | ···················· | ······T············· | ···················· |
| Q | 3667 hCoV-19/England/QEUH-A4C930/2020\|EPI_ISL_628579\|20 | ···················· | ·A·······A·········· | ···················· | ···················· | ···················· | ···················· |
| Q | 3668 hCoV-19/England/QEUH-A4C94F/2020\|EPI_ISL_628580\|20 | ···················· | ···················· | ···················· | ···················· | ···················· | ···················· |
| Q | 3669 hCoV-19/England/QEUH-A4C95E/2020\|EPI_ISL_628581\|20 | ···················· | ···················· | ···················· | ···················· | ···················· | ···················· |
| Q | 3670 hCoV-19/Scotland/QEUH-A4C98B/2020\|EPI_ISL_628582\|2 | ···················· | ···················· | ···················· | ···················· | ···················· | ···················· |
| Q | 3671 hCoV-19/Scotland/QEUH-A4C99A/2020\|EPI_ISL_628583\|2 | ···················· | ···················· | ···················· | ···················· | ···················· | ···················· |
| Q | 3672 hCoV-19/Scotland/QEUH-A4C9B8/2020\|EPI_ISL_628584\|2 | ···················· | ···················· | ···················· | ···················· | ···················· | ···················· |
| Q | 3673 hCoV-19/Scotland/QEUH-A4C9D6/2020\|EPI_ISL_628585\|2 | ···················· | ···················· | ···················· | ···················· | ···················· | ···················· |
| Q | 3674 hCoV-19/England/QEUH-A4C9F4/2020\|EPI_ISL_628586\|20 | ···················· | ···················· | ···················· | ···················· | ···················· | ···················· |
| Q | 3675 hCoV-19/England/QEUH-A4CA00/2020\|EPI_ISL_628587\|20 | ···················· | ···················· | ···················· | ···················· | ···················· | ···················· |
| Q | 3676 hCoV-19/Scotland/QEUH-A4CA2E/2020\|EPI_ISL_628588\|2 | ···················· | ···················· | ···················· | ···················· | ···················· | ···················· |
| Q | 3677 hCoV-19/Scotland/QEUH-A4CC28/2020\|EPI_ISL_628589\|2 | ···················· | ···················· | ···················· | ···················· | ···················· | ···················· |
| Q | 3678 hCoV-19/England/QEUH-A4D1EC/2020\|EPI_ISL_628590\|20 | ···················· | ···················· | ···················· | ···················· | ···················· | ···················· |
| Q | 3679 hCoV-19/Scotland/QEUH-A4D207/2020\|EPI_ISL_628591\|2 | ···················· | ···················· | ···················· | ···················· | ···················· | ···················· |
| Q | 3680 hCoV-19/Scotland/QEUH-A4D261/2020\|EPI_ISL_628592\|2 | ···················· | ···················· | ···················· | ···················· | ···················· | ···················· |
| Q | 3681 hCoV-19/England/QEUH-A4D28F/2020\|EPI_ISL_628593\|20 | ···················· | ···················· | ···················· | ···················· | ···················· | ···················· |
| Q | 3682 hCoV-19/England/QEUH-A4D313/2020\|EPI_ISL_628594\|20 | ···················· | ···················· | ···················· | ···················· | ···················· | ···················· |
| Q | 3683 hCoV-19/England/QEUH-A4D322/2020\|EPI_ISL_628595\|20 | ···················· | ···················· | ···················· | ···················· | ······T············· | ···················· |
| Q | 3684 hCoV-19/England/QEUH-A4D36E/2020\|EPI_ISL_628596\|20 | ···················· | ···················· | ···················· | ···················· | ···················· | ···················· |
| Q | 3685 hCoV-19/England/QEUH-A4D37D/2020\|EPI_ISL_628597\|20 | ···················· | ···················· | ···················· | ···················· | ···················· | ···················· |
| Q | 3686 hCoV-19/England/QEUH-A4D44D/2020\|EPI_ISL_628598\|20 | ···················· | ···················· | ···················· | ···················· | ···················· | ···················· |
| Q | 3687 hCoV-19/England/QEUH-A4D45C/2020\|EPI_ISL_628599\|20 | ···················· | ···················· | ···················· | ···················· | ···················· | ···················· |
| Q | 3688 hCoV-19/England/QEUH-AD2A87/2020\|EPI_ISL_629800\|20 | ···················· | ···················· | ···················· | ···················· | ···················· | ···················· |
| Q | 3689 hCoV-19/England/QEUH-AD18AC/2020\|EPI_ISL_629801\|20 | ···················· | ·A·················· | ···················· | ···················· | ···················· | ···················· |
| Q | 3690 hCoV-19/England/QEUH-AD1D8F/2020\|EPI_ISL_629802\|20 | ···················· | ···················· | ···················· | ···················· | ···················· | ···················· |
| Q | 3691 hCoV-19/England/QEUH-AD04C7/2020\|EPI_ISL_629803\|20 | ···················· | ···················· | ···················· | ···················· | ···················· | ···················· |
| Q | 3692 hCoV-19/England/QEUH-AD24F2/2020\|EPI_ISL_629804\|20 | ···················· | ···················· | ···················· | ···················· | ···················· | ···················· |
| Q | 3693 hCoV-19/England/QEUH-AD2340/2020\|EPI_ISL_629805\|20 | ···················· | ···················· | ···················· | ···················· | ···················· | ···················· |
| Q | 3694 hCoV-19/England/QEUH-AD1DBC/2020\|EPI_ISL_629806\|20 | ···················· | ···················· | ···················· | ···················· | ···················· | ···················· |
| Q | 3695 hCoV-19/England/QEUH-AD03E8/2020\|EPI_ISL_629829\|20 | ···················· | ·A·················· | ···················· | ···················· | ···················· | ···A················ |
| Q | 3696 hCoV-19/England/QEUH-AD2F3D/2020\|EPI_ISL_629830\|20 | ···················· | ···················· | ···················· | ···················· | ···················· | ···················· |

Q 3697 hCoV-19/England/QEUH-AD09B9/2020|EPI_ISL_629831|20 ···················· ···················· ···················· ···················· ···················· ····················
Q 3698 hCoV-19/England/QEUH-AD1806/2020|EPI_ISL_629832|20 ···················· ···················· ···················· ···················· ···················· ····················
Q 3699 hCoV-19/England/QEUH-AD1EF5/2020|EPI_ISL_629834|20 ···················· ···················· ···················· ···················· ···················· ····················
Q 3700 hCoV-19/England/QEUH-AD25C2/2020|EPI_ISL_629835|20 ···················· ···················· ···················· ···················· ···················· ····················
Q 3701 hCoV-19/England/QEUH-AD0351/2020|EPI_ISL_629836|20 ···················· ···················· ···················· ···················· ···················· ····················
Q 3702 hCoV-19/England/QEUH-AD2692/2020|EPI_ISL_629837|20 ···················· ···················· ···················· ···················· ···················· ····················
Q 3703 hCoV-19/England/QEUH-AD0F11/2020|EPI_ISL_629838|20 ···················· ···················· ···················· ···················· ···················· ····················
Q 3704 hCoV-19/England/QEUH-AD2762/2020|EPI_ISL_629840|20 ···················· ···················· ···················· ···················· ···················· ····················
Q 3705 hCoV-19/England/QEUH-AD2A4B/2020|EPI_ISL_629841|20 ···················· ···················· ···················· ···················· ···················· ····················
Q 3706 hCoV-19/England/QEUH-AD34E2/2020|EPI_ISL_629842|20 ···················· ···················· ···················· ···················· ···················· ····················
Q 3707 hCoV-19/England/QEUH-AD2E6D/2020|EPI_ISL_629843|20 ···················· ···················· ···················· ···················· ···················· ····················
Q 3708 hCoV-19/England/QEUH-AD25B3/2020|EPI_ISL_629844|20 ···················· ···················· ···················· ···················· ···················· ····················
Q 3709 hCoV-19/England/QEUH-AD632E/2020|EPI_ISL_629847|20 ···················· ···················· ···················· ···················· ···················· ····················
Q 3710 hCoV-19/England/QEUH-AD0EF6/2020|EPI_ISL_629848|20 ···················· ·A·················· ···················· ···················· ···················· ····················
Q 3711 hCoV-19/England/QEUH-AD04B8/2020|EPI_ISL_629865|20 ···················· ···················· ···················· ···················· ···················· ····················
Q 3712 hCoV-19/England/QEUH-AD3AD1/2020|EPI_ISL_629867|20 ···················· ···················· ···················· ···················· ···················· ····················
Q 3713 hCoV-19/England/QEUH-AD0667/2020|EPI_ISL_629868|20 ···················· ·A·················· ···················· ···················· ···················· ··A·················
Q 3714 hCoV-19/England/QEUH-AD045E/2020|EPI_ISL_629869|20 ···················· ···················· ···················· ···················· ···················· ····················
Q 3715 hCoV-19/England/QEUH-AD18BB/2020|EPI_ISL_629891|20 ···················· ···················· ···················· ···················· ···················· ····················
Q 3716 hCoV-19/England/QEUH-AD1C46/2020|EPI_ISL_629892|20 ···················· ···················· ···················· ···················· ···················· ····················
Q 3717 hCoV-19/England/QEUH-AD2EF4/2020|EPI_ISL_629893|20 ···················· ···················· ···················· ···················· ···················· ····················
Q 3718 hCoV-19/England/QEUH-AD2F5B/2020|EPI_ISL_629894|20 ···················· ···················· ···················· ···················· ···················· ····················
Q 3719 hCoV-19/England/QEUH-AD3E3F/2020|EPI_ISL_629895|20 ···················· ···················· ···················· ···················· ···················· ····················
Q 3720 hCoV-19/England/QEUH-AD1772/2020|EPI_ISL_629896|20 ···················· ···················· ···················· ···················· ···················· ····················
Q 3721 hCoV-19/England/QEUH-AD2ED6/2020|EPI_ISL_629897|20 ···················· ···················· ···················· ···················· ···················· ····················
Q 3722 hCoV-19/England/QEUH-AD096E/2020|EPI_ISL_629898|20 ···················· ···················· ···················· ···················· ···················· ····················
Q 3723 hCoV-19/England/QEUH-AD2E8B/2020|EPI_ISL_629899|20 ···················· ···················· ···················· ···················· ···················· ····················
Q 3724 hCoV-19/England/ALDP-AA4C5B/2020|EPI_ISL_630810|20 ···················· ···················· ···················· ···················· ···················· ····················
Q 3725 hCoV-19/England/ALDP-AA0994/2020|EPI_ISL_630811|20 ···················· ···················· ···················· ···················· ···················· ····················
Q 3726 hCoV-19/England/ALDP-AA4E46/2020|EPI_ISL_630812|20 ···················· ···················· ···················· ···················· ···················· ····················
Q 3727 hCoV-19/England/QEUH-AAE231/2020|EPI_ISL_630813|20 ···················· ···················· ···················· ···················· ···················· ····················
Q 3728 hCoV-19/England/ALDP-AA2701/2020|EPI_ISL_630814|20 ···················· ···················· ···················· ···················· ···················· ····················
Q 3729 hCoV-19/England/QEUH-AAE671/2020|EPI_ISL_630815|20 ···················· ·A·················· ···················· ···················· ···················· ····················
Q 3730 hCoV-19/England/ALDP-AA4DB2/2020|EPI_ISL_630816|20 ···················· ···················· ···················· ···················· ···················· ····················
Q 3731 hCoV-19/England/ALDP-AA26D7/2020|EPI_ISL_630817|20 ···················· ···················· ···················· ···················· ···················· ····················
Q 3732 hCoV-19/England/ALDP-AA3982/2020|EPI_ISL_630818|20 ···················· ···················· ···················· ···················· ···················· ····················
Q 3733 hCoV-19/England/ALDP-AA1BAC/2020|EPI_ISL_630819|20 ···················· ···················· ···················· ···················· ···················· ····················
Q 3734 hCoV-19/England/ALDP-AA4FAD/2020|EPI_ISL_630820|20 ···················· ···················· ···················· ···················· ···················· ····················
Q 3735 hCoV-19/England/ALDP-AA309C/2020|EPI_ISL_630821|20 ···················· ·A·················· ···················· ···················· ···················· ··A·················
Q 3736 hCoV-19/England/ALDP-AA4FDA/2020|EPI_ISL_630822|20 ···················· ···················· ···················· ···················· ···················· ····················
Q 3737 hCoV-19/England/ALDP-AA3FF9/2020|EPI_ISL_630825|20 ···················· ···················· ···················· ···················· ···················· ····················
Q 3738 hCoV-19/England/ALDP-AA2552/2020|EPI_ISL_630826|20 ···················· ·A·················· ···················· ···················· ···················· ··A·················
Q 3739 hCoV-19/England/QEUH-AAE705/2020|EPI_ISL_630827|20 ···················· ···················· ···················· ···················· ···················· ····················
Q 3740 hCoV-19/England/ALDP-AA56F2/2020|EPI_ISL_630828|20 ···················· ·A·················· ···················· ···················· ···················· ··A·················
Q 3741 hCoV-19/England/ALDP-AA4F34/2020|EPI_ISL_630829|20 ···················· ···················· ···················· ···················· ···················· ····················
Q 3742 hCoV-19/England/ALDP-AA268C/2020|EPI_ISL_630841|20 ···················· ···················· ···················· ···················· ···················· ····················
Q 3743 hCoV-19/England/ALDP-AA26E6/2020|EPI_ISL_630842|20 ···················· ···················· ···················· ···················· ···················· ····················
Q 3744 hCoV-19/England/ALDP-AA2929/2020|EPI_ISL_630843|20 ···················· ···················· ···················· ···················· ···················· ····················
Q 3745 hCoV-19/England/ALDP-AA4D94/2020|EPI_ISL_630844|20 ···················· ···················· ···················· ···················· ···················· ····················
Q 3746 hCoV-19/England/ALDP-AA4E55/2020|EPI_ISL_630845|20 ···················· ···················· ···················· ···················· ···················· ····················
Q 3747 hCoV-19/England/ALDP-AA4E19/2020|EPI_ISL_630847|20 ···················· ···················· ···················· ···················· ···················· ····················
Q 3748 hCoV-19/England/ALDP-AA38FE/2020|EPI_ISL_630848|20 ···················· ···················· ···················· ···················· ···················· ····················
Q 3749 hCoV-19/England/QEUH-AAE028/2020|EPI_ISL_630851|20 ···················· ···················· ···················· ···················· ···················· ····················
Q 3750 hCoV-19/England/ALDP-AA1C03/2020|EPI_ISL_630874|20 ···················· ···················· ···················· ···················· ···················· ····················
Q 3751 hCoV-19/England/QEUH-AAE134/2020|EPI_ISL_630875|20 ···················· ···················· ···················· ···················· ···················· ····················
Q 3752 hCoV-19/England/ALDP-AA4E91/2020|EPI_ISL_630877|20 ···················· ···················· ···················· ···················· ···················· ····················
Q 3753 hCoV-19/England/ALDP-AA2A53/2020|EPI_ISL_630878|20 ···················· ···················· ···················· ···················· ···················· ····················
Q 3754 hCoV-19/England/ALDP-AA4F9E/2020|EPI_ISL_630879|20 ···················· ···················· ···················· ···················· ···················· ····················
Q 3755 hCoV-19/England/ALDP-AA4E64/2020|EPI_ISL_630880|20 ···················· ···················· ···················· ···················· ···················· ····················
Q 3756 hCoV-19/England/ALDP-AA31D5/2020|EPI_ISL_630881|20 ···················· ···················· ···················· ···················· ···················· ····················
Q 3757 hCoV-19/England/ALDP-AA5021/2020|EPI_ISL_630882|20 ···················· ···················· ···················· ···················· ···················· ····················
Q 3758 hCoV-19/England/QEUH-AAF2A9/2020|EPI_ISL_630883|20 ···················· ···················· ···················· ···················· ···················· ····················
Q 3759 hCoV-19/England/ALDP-AA2992/2020|EPI_ISL_630884|20 ···················· ···················· ···················· ···················· ···················· ····················
Q 3760 hCoV-19/England/ALDP-AA2640/2020|EPI_ISL_630885|20 ···················· ···················· ···················· ···················· ···················· ····················
Q 3761 hCoV-19/England/ALDP-AA30D8/2020|EPI_ISL_630886|20 ···················· ···················· ···················· ···················· ···················· ····················
Q 3762 hCoV-19/England/ALDP-AA2FBE/2020|EPI_ISL_630887|20 ···················· ·A·················· ···················· ···················· ···················· ··A·················
Q 3763 hCoV-19/England/QEUH-AAE78D/2020|EPI_ISL_630888|20 ···················· ···················· ···················· ···················· ···················· ····················
Q 3764 hCoV-19/England/ALDP-AA313F/2020|EPI_ISL_630889|20 ···················· ·A·················· ···················· ···················· ···················· ····················
Q 3765 hCoV-19/England/ALDP-AA3005/2020|EPI_ISL_630890|20 ···················· ···················· ···················· ···················· ···················· ····················
Q 3766 hCoV-19/England/ALDP-AA2983/2020|EPI_ISL_630892|20 ···················· ···················· ···················· ···················· ···················· ····················
Q 3767 hCoV-19/England/ALDP-AA3973/2020|EPI_ISL_630893|20 ···················· ···················· ···················· ···················· ···················· ····················
Q 3768 hCoV-19/England/ALDP-AA570E/2020|EPI_ISL_630896|20 ···················· ···················· ···················· ···················· ···················· ····················
Q 3769 hCoV-19/England/ALDP-AA2604/2020|EPI_ISL_630897|20 ···················· ···················· ···················· ···················· ···················· ····················
Q 3770 hCoV-19/England/ALDP-AA1BBB/2020|EPI_ISL_630898|20 ···················· ···················· ···················· ···················· ···················· ····················
Q 3771 hCoV-19/England/ALDP-AA4EBF/2020|EPI_ISL_630899|20 ···················· ···················· ···················· ···················· ···················· ····················
Q 3772 hCoV-19/USA/NY-NYCPHL-001010/2020|EPI_ISL_633030|2 ···················· ···················· ···················· ···················· ···················· ····················
Q 3773 hCoV-19/USA/NY-NYCPHL-001011/2020|EPI_ISL_633031|2 ···················· ·····A·············· ···················· ···················· ···················· ····················
Q 3774 hCoV-19/USA/NY-NYCPHL-001013/2020|EPI_ISL_633032|2 ···················· ·A·················· ···················· ···················· ···················· ····················
Q 3775 hCoV-19/USA/NY-NYCPHL-001014/2020|EPI_ISL_633033|2 ···················· ···················· ···················· ···················· ···················· ····················
Q 3776 hCoV-19/USA/NY-NYCPHL-001015/2020|EPI_ISL_633034|2 ···················· ···················· ···················· ···················· ···················· ····················

| | | | | | | | |
|---|---|---|---|---|---|---|---|
| Q | 3777 hCoV-19/USA/NY-NYCPHL-001016/2020\|EPI_ISL_633035\|2 | ···················· | ·A·················· | ···················· | ···················· | ···················· | ···················· |
| Q | 3778 hCoV-19/USA/NY-NYCPHL-001018/2020\|EPI_ISL_633036\|2 | ···················· | ···················· | ···················· | ···················· | ···················· | ···················· |
| Q | 3779 hCoV-19/USA/NY-NYCPHL-001019/2020\|EPI_ISL_633037\|2 | ···················· | ·A·················· | ···················· | ···················· | ···················· | ···················· |
| Q | 3780 hCoV-19/USA/NY-NYCPHL-001020/2020\|EPI_ISL_633038\|2 | ···················· | ···················· | ···················· | ···················· | ···················· | ···················· |
| Q | 3781 hCoV-19/USA/NY-NYCPHL-001021/2020\|EPI_ISL_633039\|2 | ···················· | ···················· | ···················· | ···················· | ···················· | ···················· |
| Q | 3782 hCoV-19/USA/NY-NYCPHL-001024/2020\|EPI_ISL_633041\|2 | ···················· | ···················· | ···················· | ···················· | ···················· | ···················· |
| Q | 3783 hCoV-19/USA/NY-NYCPHL-001025/2020\|EPI_ISL_633042\|2 | ···················· | ···················· | ···················· | ···················· | ···················· | ···················· |
| Q | 3784 hCoV-19/USA/NY-NYCPHL-001028/2020\|EPI_ISL_633044\|2 | ···················· | ···················· | ···················· | ···················· | ···················· | ···················· |
| Q | 3785 hCoV-19/USA/NY-NYCPHL-001029/2020\|EPI_ISL_633045\|2 | ···················· | ·A·················· | ···················· | ···················· | ···················· | ···················· |
| Q | 3786 hCoV-19/USA/NY-NYCPHL-001043/2020\|EPI_ISL_633055\|2 | ···················· | ···················· | ···················· | ···················· | ···················· | ···················· |
| Q | 3787 hCoV-19/USA/NY-NYCPHL-001047/2020\|EPI_ISL_633059\|2 | ···················· | ···················· | ···················· | ···················· | ···················· | ···················· |
| Q | 3788 hCoV-19/USA/NY-NYCPHL-001051/2020\|EPI_ISL_633062\|2 | ···················· | ·A·················· | ···················· | ···················· | ···················· | ···················· |
| Q | 3789 hCoV-19/USA/NY-NYCPHL-001061/2020\|EPI_ISL_633067\|2 | ···················· | ·A·················· | ···················· | ···················· | ···················· | ···················· |
| Q | 3790 hCoV-19/USA/MD-MDH-0314/2020\|EPI_ISL_633090\|2020-1 | ···················· | ···················· | ···················· | ···················· | ···················· | ···················· |
| Q | 3791 hCoV-19/USA/MD-MDH-0316/2020\|EPI_ISL_633092\|2020-1 | ···················· | ·A·················· | ···················· | ···················· | ···················· | ···················· |
| Q | 3792 hCoV-19/USA/MD-MDH-0317/2020\|EPI_ISL_633093\|2020-1 | ···················· | ·A·················· | ···················· | ···················· | ···················· | ···················· |
| Q | 3793 hCoV-19/USA/MD-MDH-0319/2020\|EPI_ISL_633094\|2020-1 | ···················· | ······A············· | ···················· | ···················· | ···················· | ···················· |
| Q | 3794 hCoV-19/USA/MD-MDH-0320/2020\|EPI_ISL_633095\|2020-1 | ···················· | ···················· | ···················· | ···················· | ···················· | ···················· |
| Q | 3795 hCoV-19/USA/MD-MDH-0321/2020\|EPI_ISL_633096\|2020-1 | ···················· | ···················· | ···················· | ···················· | ···················· | ···················· |
| Q | 3796 hCoV-19/USA/MD-MDH-0322/2020\|EPI_ISL_633097\|2020-1 | ···················· | ············C······· | ···················· | ···················· | ···················· | ···················· |
| Q | 3797 hCoV-19/USA/MD-MDH-0323/2020\|EPI_ISL_633098\|2020-1 | ···················· | ·A·················· | ···················· | ···················· | ···················· | ···················· |
| Q | 3798 hCoV-19/USA/MD-MDH-0324/2020\|EPI_ISL_633099\|2020-1 | ···················· | ···················· | ···················· | ···················· | ···················· | ···················· |
| Q | 3799 hCoV-19/England/MILK-ACD65E/2020\|EPI_ISL_634360\|20 | ···················· | ···················· | ···················· | ···················· | ···················· | ···················· |
| Q | 3800 hCoV-19/England/MILK-ACCB05/2020\|EPI_ISL_634361\|20 | ···················· | ···················· | ···················· | ···················· | ···················· | ···················· |
| Q | 3801 hCoV-19/England/MILK-ACC428/2020\|EPI_ISL_634362\|20 | ···················· | ···················· | ···················· | ···················· | ···················· | ···················· |
| Q | 3802 hCoV-19/England/MILK-AC786D/2020\|EPI_ISL_634364\|20 | ···················· | ···················· | ···················· | ···················· | ···················· | ···················· |
| Q | 3803 hCoV-19/England/MILK-ACD612/2020\|EPI_ISL_634365\|20 | ···················· | ···················· | ···················· | ···················· | ···················· | ···················· |
| Q | 3804 hCoV-19/England/MILK-ACD3EE/2020\|EPI_ISL_634367\|20 | ···················· | ···················· | ···················· | ···················· | ···················· | ···················· |
| Q | 3805 hCoV-19/England/MILK-ACCF18/2020\|EPI_ISL_634368\|20 | ···················· | ···················· | ···················· | ···················· | ···················· | ···················· |
| Q | 3806 hCoV-19/England/MILK-AC7997/2020\|EPI_ISL_634369\|20 | ···················· | ···················· | ···················· | ···················· | ···················· | ···················· |
| Q | 3807 hCoV-19/England/MILK-AC7B82/2020\|EPI_ISL_634370\|20 | ···················· | ···················· | ···················· | ···················· | ······T············· | ···················· |
| Q | 3808 hCoV-19/England/MILK-ACD64F/2020\|EPI_ISL_634371\|20 | ···················· | ···················· | ···················· | ···················· | ···················· | ···················· |
| Q | 3809 hCoV-19/England/MILK-AC8161/2020\|EPI_ISL_634372\|20 | ···················· | ···················· | ···················· | ···················· | ······T············· | ···················· |
| Q | 3810 hCoV-19/England/MILK-AC7AA3/2020\|EPI_ISL_634373\|20 | ···················· | ···················· | ···················· | ···················· | ···················· | ···················· |
| Q | 3811 hCoV-19/England/MILK-AC75C0/2020\|EPI_ISL_634374\|20 | ···················· | ···················· | ···················· | ···················· | ···················· | ···················· |
| Q | 3812 hCoV-19/England/MILK-AC8143/2020\|EPI_ISL_634375\|20 | ···················· | ···················· | ···················· | ···················· | ···················· | ···················· |
| Q | 3813 hCoV-19/England/MILK-ACC464/2020\|EPI_ISL_634378\|20 | ···················· | ···················· | ···················· | ···················· | ···················· | ···················· |
| Q | 3814 hCoV-19/England/MILK-ACD6E5/2020\|EPI_ISL_634379\|20 | ···················· | ···················· | ···················· | ···················· | ···················· | ···················· |
| Q | 3815 hCoV-19/England/MILK-AC7C34/2020\|EPI_ISL_634380\|20 | ···················· | ···················· | ···················· | ···················· | ···················· | ···················· |
| Q | 3816 hCoV-19/England/MILK-ACCAF9/2020\|EPI_ISL_634381\|20 | ···················· | ···················· | ···················· | ···················· | ···················· | ···················· |
| Q | 3817 hCoV-19/England/MILK-ACD9BF/2020\|EPI_ISL_634382\|20 | ···················· | ···················· | ···················· | ···················· | ···················· | ···················· |
| Q | 3818 hCoV-19/England/MILK-ACDA07/2020\|EPI_ISL_634383\|20 | ···················· | ·A·················· | ···················· | ···················· | ···················· | ···A················ |
| Q | 3819 hCoV-19/England/MILK-ACC91A/2020\|EPI_ISL_634386\|20 | ···················· | ···················· | ···················· | ···················· | ···················· | ···················· |
| Q | 3820 hCoV-19/England/MILK-AC80CE/2020\|EPI_ISL_634387\|20 | ···················· | ···················· | ···················· | ···················· | ···················· | ···················· |
| Q | 3821 hCoV-19/England/MILK-ACD5F7/2020\|EPI_ISL_634388\|20 | ···················· | ···················· | ···················· | ···················· | ···················· | ···················· |
| Q | 3822 hCoV-19/England/MILK-ACD83A/2020\|EPI_ISL_634389\|20 | ···················· | ···················· | ···················· | ···················· | ···················· | ···················· |
| Q | 3823 hCoV-19/England/MILK-AC80EC/2020\|EPI_ISL_634390\|20 | ···················· | ···················· | ···················· | ···················· | ···················· | ···················· |
| Q | 3824 hCoV-19/England/MILK-ACD3C0/2020\|EPI_ISL_634392\|20 | ···················· | ···················· | ···················· | ···················· | ···················· | ···················· |
| Q | 3825 hCoV-19/England/MILK-AC7AFE/2020\|EPI_ISL_634393\|20 | ···················· | ···················· | ···················· | ···················· | ····T··············· | ···················· |
| Q | 3826 hCoV-19/England/MILK-AC7E5C/2020\|EPI_ISL_634394\|20 | ···················· | ···················· | ···················· | ···················· | ···················· | ···················· |
| Q | 3827 hCoV-19/England/MILK-ACD533/2020\|EPI_ISL_634396\|20 | ···················· | ···················· | ···················· | ···················· | ···················· | ···················· |
| Q | 3828 hCoV-19/England/MILK-ACD700/2020\|EPI_ISL_634397\|20 | ···················· | ···················· | ···················· | ···················· | ···················· | ···················· |
| Q | 3829 hCoV-19/England/MILK-AC7AC1/2020\|EPI_ISL_634398\|20 | ···················· | ···················· | ···················· | ···················· | ···················· | ···················· |
| Q | 3830 hCoV-19/England/MILK-ACCEFD/2020\|EPI_ISL_634399\|20 | ···················· | ···················· | ···················· | ···················· | ···················· | ···················· |
| Q | 3831 hCoV-19/USA/CA-ALSR-2809/2020\|EPI_ISL_635690\|2020- | ···················· | ·A·················· | ···················· | ···················· | ···················· | ···················· |
| Q | 3832 hCoV-19/USA/CA-ALSR-2810/2020\|EPI_ISL_635691\|2020- | ···················· | ···················· | ···················· | ···················· | ···················· | ···················· |
| Q | 3833 hCoV-19/USA/CA-ALSR-2812/2020\|EPI_ISL_635692\|2020- | ···················· | ···················· | ···················· | ···················· | ···················· | ···················· |
| Q | 3834 hCoV-19/USA/CA-ALSR-2814/2020\|EPI_ISL_635693\|2020- | ···················· | ···················· | ···················· | ···················· | ···················· | ···················· |
| Q | 3835 hCoV-19/USA/CA-ALSR-2818/2020\|EPI_ISL_635694\|2020- | ···················· | ·A·················· | ···················· | ···················· | ···················· | ···················· |
| Q | 3836 hCoV-19/USA/CA-ALSR-2819/2020\|EPI_ISL_635695\|2020- | ···················· | ·A·················· | ···················· | ···················· | ···················· | ···················· |
| Q | 3837 hCoV-19/USA/CA-ALSR-2820/2020\|EPI_ISL_635696\|2020- | ···················· | ·A·················· | ···················· | ···················· | ···················· | ···················· |
| Q | 3838 hCoV-19/USA/CA-ALSR-2822/2020\|EPI_ISL_635697\|2020- | ···················· | ·A·················· | ···················· | ···················· | ···················· | ···················· |
| Q | 3839 hCoV-19/USA/CA-ALSR-2823/2020\|EPI_ISL_635698\|2020- | ···················· | ·A·················· | ···················· | ···················· | ···················· | ···················· |
| Q | 3840 hCoV-19/USA/CA-ALSR-2825/2020\|EPI_ISL_635699\|2020- | ···················· | ···················· | ···················· | ···················· | ···················· | ···················· |
| Q | 3841 hCoV-19/Wales/CVR4765/2020\|EPI_ISL_627208\|2020-09- | ···················· | ···················· | ···················· | ···················· | ···················· | ···················· |
| Q | 3842 hCoV-19/England/EXET-138A4C/2020\|EPI_ISL_627210\|20 | ···················· | ···················· | ···················· | ···················· | ···················· | ···················· |
| Q | 3843 hCoV-19/England/LOND-DC38C/2020\|EPI_ISL_627211\|202 | ···················· | ···················· | ···················· | ···················· | ···················· | ···················· |
| Q | 3844 hCoV-19/England/MILK-A18EC5/2020\|EPI_ISL_627212\|20 | ···················· | ···················· | ···················· | ···················· | ···················· | ···················· |
| Q | 3845 hCoV-19/England/NORW-F15E5/2020\|EPI_ISL_627213\|202 | ···················· | ···················· | ···················· | ···················· | ···················· | ···················· |
| Q | 3846 hCoV-19/England/NORW-F1B4D/2020\|EPI_ISL_627214\|202 | ···················· | ···················· | ···················· | ···················· | ···················· | ···················· |
| Q | 3847 hCoV-19/England/NOTT-11907C/2020\|EPI_ISL_627215\|20 | ···················· | ···················· | ···················· | ···················· | ···················· | ···················· |
| Q | 3848 hCoV-19/England/NOTT-11972C/2020\|EPI_ISL_627216\|20 | ···················· | ···················· | ···················· | ···················· | ···················· | ···················· |
| Q | 3849 hCoV-19/Wales/PHWC-482D29/2020\|EPI_ISL_627217\|2020 | ···················· | ···················· | ···················· | ···················· | ···················· | ···················· |
| Q | 3850 hCoV-19/Wales/PHWC-482E71/2020\|EPI_ISL_627218\|2020 | ···················· | ···················· | ···················· | ···················· | ···················· | ···················· |
| Q | 3851 hCoV-19/Wales/PHWC-4834C9/2020\|EPI_ISL_627219\|2020 | ···················· | ···················· | ···················· | ···················· | ···················· | ···················· |
| Q | 3852 hCoV-19/Wales/PHWC-483511/2020\|EPI_ISL_627220\|2020 | ···················· | ···················· | ···················· | ···················· | ···················· | ···················· |
| Q | 3853 hCoV-19/Wales/PHWC-483836/2020\|EPI_ISL_627221\|2020 | ···················· | ···················· | ···················· | ···················· | ···················· | ···················· |
| Q | 3854 hCoV-19/Wales/PHWC-483942/2020\|EPI_ISL_627222\|2020 | ···················· | ···················· | ···················· | ···················· | ···················· | ···················· |
| Q | 3855 hCoV-19/Wales/PHWC-4839AC/2020\|EPI_ISL_627223\|2020 | ···················· | ···················· | ···················· | ···················· | ···················· | ···················· |
| Q | 3856 hCoV-19/Wales/PHWC-483B5B/2020\|EPI_ISL_627224\|2020 | ···················· | ···················· | ···················· | ···················· | ···················· | ···················· |

Q 3858 hCoV-19/Wales/PHWC-48411C/2020|EPI_ISL_627226|2020 ···················· ···················· ···················· ···················· ···················· ····················
Q 3859 hCoV-19/Wales/PHWC-484158/2020|EPI_ISL_627227|2020 ···················· ···················· ···················· ···················· ···················· ····················
Q 3861 hCoV-19/Wales/PHWC-48445F/2020|EPI_ISL_627229|2020 ···················· ···················· ···················· ···················· ···················· ····················
Q 3862 hCoV-19/Scotland/QEUH-A4B7CE/2020|EPI_ISL_628480|2 ···················· ···················· ···················· ···················· ···················· ····················
Q 3863 hCoV-19/Scotland/QEUH-A4B7EC/2020|EPI_ISL_628481|2 ···················· ···················· ···················· ···················· ···················· ····················
Q 3864 hCoV-19/Scotland/QEUH-A4B807/2020|EPI_ISL_628482|2 ···················· ···················· ···················· ···················· ···················· ····················
Q 3865 hCoV-19/Scotland/QEUH-A4B825/2020|EPI_ISL_628483|2 ···················· ···················· ···················· ···················· ···················· ····················
Q 3866 hCoV-19/Scotland/QEUH-A4B834/2020|EPI_ISL_628484|2 ···················· ···················· ···················· ···················· ···················· ····················
Q 3867 hCoV-19/Scotland/QEUH-A4B861/2020|EPI_ISL_628485|2 ···················· ···················· ···················· ···················· ···················· ····················
Q 3868 hCoV-19/Scotland/QEUH-A4B88F/2020|EPI_ISL_628486|2 ···················· ···················· ···················· ···················· ···················· ····················
Q 3869 hCoV-19/Scotland/QEUH-A4B89E/2020|EPI_ISL_628487|2 ···················· ···················· ···················· ···················· ···················· ····················
Q 3870 hCoV-19/Scotland/QEUH-A4B8BC/2020|EPI_ISL_628488|2 ···················· ···················· ···················· ···················· ···················· ····················
Q 3871 hCoV-19/Scotland/QEUH-A4B8CB/2020|EPI_ISL_628489|2 ···················· ···················· ···················· ···················· ···················· ····················
Q 3872 hCoV-19/Scotland/QEUH-A4B8E9/2020|EPI_ISL_628490|2 ···················· ···················· ···················· ···················· ···················· ····················
Q 3873 hCoV-19/Scotland/QEUH-A4B8F8/2020|EPI_ISL_628491|2 ···················· ···················· ···················· ···················· ···················· ····················
Q 3874 hCoV-19/Scotland/QEUH-A4B904/2020|EPI_ISL_628492|2 ···················· ···················· ···················· ···················· ···················· ····················
Q 3875 hCoV-19/Scotland/QEUH-A4B913/2020|EPI_ISL_628493|2 ···················· ···················· ···················· ···················· ···················· ····················
Q 3876 hCoV-19/Scotland/QEUH-A4B922/2020|EPI_ISL_628494|2 ···················· ···················· ···················· ···················· ···················· ····················
Q 3877 hCoV-19/Scotland/QEUH-A4B940/2020|EPI_ISL_628495|2 ···················· ···················· ···················· ···················· ···················· ····················
Q 3878 hCoV-19/Scotland/QEUH-A4B98C/2020|EPI_ISL_628496|2 ···················· ···················· ···················· ···················· ···················· ····················
Q 3879 hCoV-19/Scotland/QEUH-A4B99B/2020|EPI_ISL_628497|2 ···················· ···················· ···················· ···················· ···················· ····················
Q 3880 hCoV-19/Scotland/QEUH-A4B9AA/2020|EPI_ISL_628498|2 ···················· ···················· ···················· ···················· ···················· ····················
Q 3881 hCoV-19/Scotland/QEUH-A4B9B9/2020|EPI_ISL_628499|2 ···················· ···················· ···················· ···················· ···················· ····················
Q 3882 hCoV-19/England/QEUH-A4BC83/2020|EPI_ISL_628520|20 ···················· ···················· ···················· ···················· ···················· ····················
Q 3883 hCoV-19/England/QEUH-A4BCA1/2020|EPI_ISL_628521|20 ···················· ···················· ···················· ···················· ···················· ····················
Q 3884 hCoV-19/England/QEUH-A4BCCF/2020|EPI_ISL_628522|20 ···················· ···················· ···················· ···················· ···················· ····················
Q 3885 hCoV-19/England/QEUH-A4BD08/2020|EPI_ISL_628523|20 ···················· ···················· ···················· ···················· ···················· ····················
Q 3886 hCoV-19/England/QEUH-A4BD17/2020|EPI_ISL_628524|20 ···················· ···················· ···················· ···················· ···················· ····················
Q 3887 hCoV-19/England/QEUH-A4BD26/2020|EPI_ISL_628525|20 ···················· ···················· ···················· ···················· ···················· ····················
Q 3888 hCoV-19/England/QEUH-A4BD62/2020|EPI_ISL_628526|20 ···················· ···················· ···················· ···················· ···················· ····················
Q 3889 hCoV-19/England/QEUH-A4BD71/2020|EPI_ISL_628527|20 ···················· ···················· ···················· ···················· ···················· ····················
Q 3890 hCoV-19/England/QEUH-A4BD80/2020|EPI_ISL_628528|20 ···················· ···················· ···················· ···················· ···················· ····················
Q 3891 hCoV-19/England/QEUH-A4BDCC/2020|EPI_ISL_628529|20 ···················· ···················· ···················· ···················· ···················· ····················
Q 3892 hCoV-19/England/QEUH-A4BE41/2020|EPI_ISL_628530|20 ···················· ···················· ···················· ···················· ···················· ····················
Q 3893 hCoV-19/Scotland/QEUH-A4BE6F/2020|EPI_ISL_628531|2 ···················· ···················· ···················· ···················· ···················· ····················
Q 3894 hCoV-19/England/QEUH-A4BF5D/2020|EPI_ISL_628532|20 ···················· ···················· ···················· ···················· ···················· ····················
Q 3895 hCoV-19/England/QEUH-A4BF6C/2020|EPI_ISL_628533|20 ···················· ···················· ···················· ···················· ···················· ····················
Q 3896 hCoV-19/England/QEUH-A4C059/2020|EPI_ISL_628540|20 ···················· ···················· ···················· ···················· ···················· ····················
Q 3897 hCoV-19/England/QEUH-A4C086/2020|EPI_ISL_628541|20 ···················· ···················· ···················· ···················· ···················· ····················
Q 3898 hCoV-19/England/QEUH-A4C0B3/2020|EPI_ISL_628542|20 ···················· ···················· ···················· ···················· ···················· ····················
Q 3899 hCoV-19/England/QEUH-A4C0C2/2020|EPI_ISL_628543|20 ···················· ···················· ···················· ···················· ···················· ····················
Q 3900 hCoV-19/England/QEUH-A4C0FF/2020|EPI_ISL_628544|20 ···················· ···················· ···················· ···················· ···················· ····················
Q 3901 hCoV-19/England/QEUH-A4C138/2020|EPI_ISL_628545|20 ···················· ···················· ···················· ···················· ···················· ····················
Q 3902 hCoV-19/England/QEUH-A4C147/2020|EPI_ISL_628546|20 ···················· ···················· ···················· ···················· ···················· ····················
Q 3903 hCoV-19/England/QEUH-A4C183/2020|EPI_ISL_628547|20 ···················· ···················· ···················· ···················· ····T··············· ····················
Q 3904 hCoV-19/England/QEUH-A4C192/2020|EPI_ISL_628548|20 ···················· ···················· ···················· ···················· ···················· ····················
Q 3905 hCoV-19/Scotland/QEUH-A4C1B0/2020|EPI_ISL_628549|2 ···················· ···················· ···················· ···················· ···················· ····················
Q 3906 hCoV-19/Scotland/QEUH-A4C1ED/2020|EPI_ISL_628550|2 ···················· ···················· ···················· ···················· ···················· ····················
Q 3907 hCoV-19/Scotland/QEUH-A4C1FC/2020|EPI_ISL_628551|2 ···················· ···················· ···················· ···················· ···················· ····················
Q 3908 hCoV-19/Scotland/QEUH-A4C217/2020|EPI_ISL_628552|2 ···················· ···················· ···················· ···················· ···················· ····················
Q 3909 hCoV-19/Scotland/QEUH-A4C235/2020|EPI_ISL_628553|2 ···················· ···················· ···················· ···················· ···················· ····················
Q 3910 hCoV-19/Scotland/QEUH-A4C244/2020|EPI_ISL_628554|2 ···················· ···················· ···················· ···················· ···················· ····················
Q 3911 hCoV-19/Scotland/QEUH-A4C253/2020|EPI_ISL_628555|2 ···················· ···················· ···················· ···················· ···················· ····················
Q 3912 hCoV-19/Scotland/QEUH-A4C262/2020|EPI_ISL_628556|2 ···················· ···················· ···················· ···················· ···················· ····················
Q 3913 hCoV-19/Scotland/QEUH-A4C280/2020|EPI_ISL_628557|2 ···················· ···················· ···················· ···················· ···················· ····················
Q 3914 hCoV-19/Scotland/QEUH-A4C29F/2020|EPI_ISL_628558|2 ···················· ···················· ···················· ···················· ···················· ····················
Q 3915 hCoV-19/England/QEUH-A4C2DB/2020|EPI_ISL_628559|20 ···················· ···················· ···················· ···················· ···················· ····················
Q 3916 hCoV-19/England/QEUH-AD3AB3/2020|EPI_ISL_629850|20 ···················· ···················· ···················· ···················· ···················· ····················
Q 3917 hCoV-19/England/QEUH-AD1842/2020|EPI_ISL_629851|20 ···················· ···················· ···················· ···················· ···················· ····················
Q 3918 hCoV-19/England/QEUH-AD2EB8/2020|EPI_ISL_629852|20 ···················· ···················· ···················· ···················· ···················· ····················
Q 3919 hCoV-19/England/QEUH-AD3EC6/2020|EPI_ISL_629853|20 ···················· ···················· ···················· ···················· ···················· ····················
Q 3920 hCoV-19/England/QEUH-AD2234/2020|EPI_ISL_629854|20 ···················· ···················· ···················· ···················· ···················· ····················
Q 3921 hCoV-19/England/QEUH-AD3E4E/2020|EPI_ISL_629855|20 ···················· ···················· ···················· ···················· ···················· ····················
Q 3922 hCoV-19/England/QEUH-AD22DA/2020|EPI_ISL_629856|20 ···················· ···················· ···················· ···················· ···················· ····················
Q 3923 hCoV-19/England/QEUH-AD295D/2020|EPI_ISL_629857|20 ···················· ···················· ···················· ···················· ···················· ····················
Q 3924 hCoV-19/England/QEUH-AD2331/2020|EPI_ISL_629858|20 ···················· ···················· ···················· ···················· ···················· ····················
Q 3925 hCoV-19/England/QEUH-AD2D60/2020|EPI_ISL_629859|20 ···················· ···················· ···················· ···················· ···················· ····················
Q 3926 hCoV-19/England/QEUH-AD252C/2020|EPI_ISL_629861|20 ···················· ···················· ···················· ···················· ···················· ····················
Q 3927 hCoV-19/England/QEUH-AD2586/2020|EPI_ISL_629862|20 ···················· ···················· ···················· ···················· ···················· ····················
Q 3928 hCoV-19/England/QEUH-AD1745/2020|EPI_ISL_629863|20 ···················· ···················· ···················· ···················· ···················· ····················
Q 3929 hCoV-19/England/QEUH-AD2577/2020|EPI_ISL_629864|20 ···················· ···················· ···················· ···················· ···················· ····················
Q 3930 hCoV-19/England/QEUH-AD1851/2020|EPI_ISL_629870|20 ···················· ···················· ···················· ···················· ···················· ····················
Q 3931 hCoV-19/England/QEUH-AD3A68/2020|EPI_ISL_629871|20 ···················· ···················· ···················· ···················· ···················· ····················
Q 3932 hCoV-19/England/QEUH-AD39F2/2020|EPI_ISL_629872|20 ···················· ···················· ···················· ···················· ···················· ····················
Q 3933 hCoV-19/England/QEUH-AD0658/2020|EPI_ISL_629873|20 ···················· ···················· ···················· ···················· ···················· ····················
Q 3934 hCoV-19/England/QEUH-AD2E5E/2020|EPI_ISL_629874|20 ···················· ···················· ···················· ···················· ···················· ····················
Q 3935 hCoV-19/England/QEUH-AD1CFB/2020|EPI_ISL_629875|20 ···················· ···················· ···················· ···················· ···················· ····················
Q 3936 hCoV-19/England/QEUH-AD343D/2020|EPI_ISL_629876|20 ···················· ···················· ···················· ···················· ···················· ····················
Q 3937 hCoV-19/England/QEUH-AD3DBA/2020|EPI_ISL_629878|20 ···················· ···················· ···················· ···················· ···················· ····················
Q 3938 hCoV-19/England/QEUH-AD2683/2020|EPI_ISL_629879|20 ···················· ···················· ···················· ···················· ···················· ····················

| | | | | | | | | |
|---|---|---|---|---|---|---|---|---|
| Q | 3939 | hCoV-19/England/QEUH-AD2DE8/2020\|EPI_ISL_629880\|20 | ···················· | ···················· | ···················· | ···················· | ···················· | ···················· |
| Q | 3940 | hCoV-19/England/QEUH-AD29F3/2020\|EPI_ISL_629884\|20 | ···················· | ·A·················· | ···················· | ···················· | ···················· | ···A················ |
| Q | 3941 | hCoV-19/England/QEUH-AD34C4/2020\|EPI_ISL_629885\|20 | ···················· | ·A·················· | ···················· | ···················· | ···················· | ···A················ |
| Q | 3942 | hCoV-19/England/QEUH-AD228F/2020\|EPI_ISL_629886\|20 | ···················· | ···················· | ···················· | ···················· | ···················· | ···················· |
| Q | 3943 | hCoV-19/England/QEUH-AD1CEC/2020\|EPI_ISL_629889\|20 | ···················· | ···················· | ···················· | ···················· | ···················· | ···················· |
| Q | 3944 | hCoV-19/England/ALDP-AA2A26/2020\|EPI_ISL_630797\|20 | ···················· | ···················· | ···················· | ···················· | ···················· | ···················· |
| Q | 3945 | hCoV-19/England/ALDP-AA4E0A/2020\|EPI_ISL_630798\|20 | ···················· | ···················· | ···················· | ···················· | ···················· | ···················· |
| Q | 3946 | hCoV-19/England/ALDP-AA272F/2020\|EPI_ISL_630799\|20 | ···················· | ···················· | ···················· | ···················· | ···················· | ···················· |
| Q | 3947 | hCoV-19/England/ALDP-AA4AF7/2020\|EPI_ISL_630830\|20 | ···················· | ···················· | ···················· | ···················· | ···················· | ···················· |
| Q | 3948 | hCoV-19/England/ALDP-AA2FCD/2020\|EPI_ISL_630831\|20 | ···················· | ·A·················· | ···················· | ···················· | ···················· | ···A················ |
| Q | 3949 | hCoV-19/England/ALDP-AA4E37/2020\|EPI_ISL_630832\|20 | ···················· | ···················· | ···················· | ···················· | ···················· | ···················· |
| Q | 3950 | hCoV-19/England/QEUH-AAE723/2020\|EPI_ISL_630833\|20 | ···················· | ···················· | ···················· | ···················· | ···················· | ···················· |
| Q | 3951 | hCoV-19/England/ALDP-AA29CF/2020\|EPI_ISL_630835\|20 | ···················· | ···················· | ···················· | ···················· | ···················· | ···················· |
| Q | 3952 | hCoV-19/England/ALDP-AA3014/2020\|EPI_ISL_630836\|20 | ···················· | ·A·················· | ···················· | ···················· | ···················· | ···A················ |
| Q | 3953 | hCoV-19/England/ALDP-AA0967/2020\|EPI_ISL_630837\|20 | ···················· | ···················· | ···················· | ···················· | ···················· | ···················· |
| Q | 3954 | hCoV-19/England/ALDP-AA573B/2020\|EPI_ISL_630840\|20 | ···················· | ···················· | ···················· | ···················· | ···················· | ···················· |
| Q | 3955 | hCoV-19/England/ALDP-AA25F8/2020\|EPI_ISL_630852\|20 | ···················· | ···················· | ···················· | ···················· | ···················· | ···················· |
| Q | 3956 | hCoV-19/England/QEUH-AAE6F9/2020\|EPI_ISL_630853\|20 | ···················· | ···················· | ···················· | ···················· | ···················· | ···················· |
| Q | 3957 | hCoV-19/England/ALDP-AA266E/2020\|EPI_ISL_630854\|20 | ···················· | ···················· | ···················· | ···················· | ···················· | ···················· |
| Q | 3958 | hCoV-19/England/ALDP-AA4D2B/2020\|EPI_ISL_630855\|20 | ···················· | ···················· | ···················· | ···················· | ···················· | ···················· |
| Q | 3959 | hCoV-19/England/ALDP-AA50F4/2020\|EPI_ISL_630856\|20 | ···················· | ···················· | ···················· | ···················· | ···················· | ···················· |
| Q | 3960 | hCoV-19/England/ALDP-AA4022/2020\|EPI_ISL_630857\|20 | ···················· | ···················· | ···················· | ···················· | ···················· | ···················· |
| Q | 3961 | hCoV-19/England/ALDP-AA4CB5/2020\|EPI_ISL_630858\|20 | ···················· | ···················· | ···················· | ···················· | ···················· | ···················· |
| Q | 3962 | hCoV-19/England/QEUH-AAE626/2020\|EPI_ISL_630859\|20 | ···················· | ···················· | ···················· | ···················· | ···················· | ···················· |
| Q | 3963 | hCoV-19/England/ALDP-AA1C4F/2020\|EPI_ISL_630860\|20 | ···················· | ···················· | ···················· | ···················· | ···················· | ···················· |
| Q | 3964 | hCoV-19/England/ALDP-AA4D67/2020\|EPI_ISL_630863\|20 | ···················· | ···················· | ···················· | ···················· | ···················· | ···················· |
| Q | 3965 | hCoV-19/England/QEUH-AAE0DD/2020\|EPI_ISL_630864\|20 | ···················· | ···················· | ···················· | ···················· | ···················· | ···················· |
| Q | 3966 | hCoV-19/England/ALDP-AA0976/2020\|EPI_ISL_630865\|20 | ···················· | ···················· | ···················· | ···················· | ···················· | ···················· |
| Q | 3967 | hCoV-19/England/ALDP-AA4040/2020\|EPI_ISL_630866\|20 | ···················· | ··············C····· | ···················· | ···················· | ···················· | ···················· |
| Q | 3968 | hCoV-19/England/ALDP-AA4AD9/2020\|EPI_ISL_630867\|20 | ···················· | ···················· | ···················· | ···················· | ···················· | ···················· |
| Q | 3969 | hCoV-19/England/ALDP-AA2A71/2020\|EPI_ISL_630868\|20 | ···················· | ···················· | ···················· | ···················· | ···················· | ···················· |
| Q | 3970 | hCoV-19/England/ALDP-AA5100/2020\|EPI_ISL_630869\|20 | ···················· | ···················· | ···················· | ···················· | ···················· | ···················· |
| Q | 3971 | hCoV-19/England/QEUH-AAE0A0/2020\|EPI_ISL_630871\|20 | ···················· | ···················· | ···················· | ···················· | ···················· | ···················· |
| Q | 3972 | hCoV-19/England/QEUH-AAE0BF/2020\|EPI_ISL_630872\|20 | ···················· | ·A·················· | ···················· | ···················· | ···················· | ···················· |
| Q | 3973 | hCoV-19/England/QEUH-AAE082/2020\|EPI_ISL_630873\|20 | ···················· | ·A·················· | ···················· | ···················· | ···················· | ···················· |
| Q | 3974 | hCoV-19/USA/NY-NYCPHL-000969/2020\|EPI_ISL_633000\|2 | ···················· | ···················· | ···················· | ···················· | ···················· | ···················· |
| Q | 3975 | hCoV-19/USA/NY-NYCPHL-000970/2020\|EPI_ISL_633001\|2 | ···················· | ···················· | ···················· | ···················· | ···················· | ···················· |
| Q | 3976 | hCoV-19/USA/NY-NYCPHL-000971/2020\|EPI_ISL_633002\|2 | ···················· | ···················· | ···················· | ···················· | ···················· | ···················· |
| Q | 3977 | hCoV-19/USA/NY-NYCPHL-000973/2020\|EPI_ISL_633003\|2 | ···················· | ···················· | ···················· | ···················· | ···················· | ···················· |
| Q | 3978 | hCoV-19/USA/NY-NYCPHL-000974/2020\|EPI_ISL_633004\|2 | ···················· | ···················· | ···················· | ···················· | ···················· | ···················· |
| Q | 3979 | hCoV-19/USA/NY-NYCPHL-000976/2020\|EPI_ISL_633005\|2 | ···················· | ············C······· | ···················· | ···················· | ···················· | ···················· |
| Q | 3980 | hCoV-19/USA/NY-NYCPHL-000978/2020\|EPI_ISL_633006\|2 | ···················· | ············C······· | ···················· | ···················· | ···················· | ···················· |
| Q | 3981 | hCoV-19/USA/NY-NYCPHL-000979/2020\|EPI_ISL_633007\|2 | ···················· | ···················· | ···················· | ···················· | ···················· | ···················· |
| Q | 3982 | hCoV-19/USA/NY-NYCPHL-000983/2020\|EPI_ISL_633010\|2 | ···················· | ·A·················· | ···················· | ···················· | ···················· | ···················· |
| Q | 3983 | hCoV-19/USA/NY-NYCPHL-000987/2020\|EPI_ISL_633013\|2 | ···················· | ···················· | ···················· | ···················· | ···················· | ···················· |
| Q | 3984 | hCoV-19/USA/NY-NYCPHL-000988/2020\|EPI_ISL_633014\|2 | ···················· | ·A·················· | ···················· | ···················· | ···················· | ···················· |
| Q | 3985 | hCoV-19/USA/NY-NYCPHL-000989/2020\|EPI_ISL_633015\|2 | ···················· | ···················· | ···················· | ···················· | ···················· | ···················· |
| Q | 3986 | hCoV-19/USA/NY-NYCPHL-000990/2020\|EPI_ISL_633016\|2 | ···················· | ·A·················· | ···················· | ···················· | ···················· | ···················· |
| Q | 3987 | hCoV-19/USA/NY-NYCPHL-000992/2020\|EPI_ISL_633017\|2 | ···················· | ···················· | ···················· | ···················· | ···················· | ···················· |
| Q | 3988 | hCoV-19/USA/NY-NYCPHL-000994/2020\|EPI_ISL_633019\|2 | ···················· | ···················· | ···················· | ···················· | ···················· | ···················· |
| Q | 3989 | hCoV-19/USA/NY-NYCPHL-001002/2020\|EPI_ISL_633025\|2 | ···················· | ···················· | ···················· | ···················· | ···················· | ···················· |
| Q | 3990 | hCoV-19/USA/NY-NYCPHL-001004/2020\|EPI_ISL_633027\|2 | ···················· | ···················· | ···················· | ···················· | ···················· | ···················· |
| Q | 3991 | hCoV-19/USA/NY-NYCPHL-001008/2020\|EPI_ISL_633029\|2 | ···················· | ···················· | ···················· | ···················· | ···················· | ···················· |
| Q | 3992 | hCoV-19/England/MILK-ACC6E6/2020\|EPI_ISL_634317\|20 | ···················· | ···················· | ···················· | ···················· | ···················· | ···················· |
| Q | 3993 | hCoV-19/England/MILK-ACC81D/2020\|EPI_ISL_634319\|20 | ···················· | ···················· | ···················· | ···················· | ···················· | ···················· |
| Q | 3994 | hCoV-19/England/MILK-ACC67D/2020\|EPI_ISL_634321\|20 | ···················· | ···················· | ···················· | ···················· | ···················· | ···················· |
| Q | 3995 | hCoV-19/England/MILK-ACD542/2020\|EPI_ISL_634322\|20 | ···················· | ···················· | ···················· | ···················· | ···················· | ···················· |
| Q | 3996 | hCoV-19/England/MILK-AC7A67/2020\|EPI_ISL_634323\|20 | ···················· | ···················· | ···················· | ···················· | ······T············· | ···················· |
| Q | 3997 | hCoV-19/England/MILK-AC75DF/2020\|EPI_ISL_634324\|20 | ···················· | ···················· | ···················· | ···················· | ···················· | ···················· |
| Q | 3998 | hCoV-19/England/MILK-ACD57F/2020\|EPI_ISL_634325\|20 | ···················· | ···················· | ···················· | ···················· | ···················· | ···················· |
| Q | 3999 | hCoV-19/England/MILK-ACC90B/2020\|EPI_ISL_634327\|20 | ···················· | ·A·················· | ···················· | ···················· | ···················· | ···A················ |
| Q | 4000 | hCoV-19/England/MILK-AC794C/2020\|EPI_ISL_634328\|20 | ···················· | ···················· | ···················· | ···················· | ···················· | ···················· |
| Q | 4001 | hCoV-19/England/MILK-ACC507/2020\|EPI_ISL_634329\|20 | ···················· | ···················· | ···················· | ···················· | ···················· | ···················· |
| Q | 4002 | hCoV-19/England/MILK-ACCBF6/2020\|EPI_ISL_634330\|20 | ···················· | ···················· | ···················· | ···················· | ···················· | ···················· |
| Q | 4003 | hCoV-19/England/MILK-AC7EA7/2020\|EPI_ISL_634331\|20 | ···················· | ·A·················· | ···················· | ···················· | ···················· | ···A················ |
| Q | 4004 | hCoV-19/England/MILK-ACD5AC/2020\|EPI_ISL_634332\|20 | ···················· | ···················· | ···················· | ···················· | ···················· | ···················· |
| Q | 4005 | hCoV-19/England/MILK-AC78C7/2020\|EPI_ISL_634334\|20 | ···················· | ···················· | ···················· | ···················· | ······T············· | ···················· |
| Q | 4006 | hCoV-19/England/MILK-AC80FB/2020\|EPI_ISL_634335\|20 | ···················· | ···················· | ···················· | ···················· | ···················· | ···················· |
| Q | 4007 | hCoV-19/England/MILK-AC8125/2020\|EPI_ISL_634336\|20 | ···················· | ···················· | ···················· | ···················· | ···················· | ···················· |
| Q | 4008 | hCoV-19/England/MILK-AC78D6/2020\|EPI_ISL_634340\|20 | ···················· | ···················· | ···················· | ···················· | ······T············· | ···················· |
| Q | 4009 | hCoV-19/England/MILK-ACC4FB/2020\|EPI_ISL_634345\|20 | ···················· | ···················· | ···················· | ···················· | ···················· | ···················· |
| Q | 4010 | hCoV-19/England/MILK-AC79C4/2020\|EPI_ISL_634346\|20 | ···················· | ···················· | ···················· | ···················· | ···················· | ···················· |
| Q | 4011 | hCoV-19/England/MILK-AC78F4/2020\|EPI_ISL_634347\|20 | ···················· | ···················· | ···················· | ···················· | ···················· | ···················· |
| Q | 4012 | hCoV-19/England/MILK-AC7B0A/2020\|EPI_ISL_634348\|20 | ···················· | ···················· | ···················· | ···················· | ···················· | ···················· |
| Q | 4013 | hCoV-19/England/MILK-AC7C9E/2020\|EPI_ISL_634349\|20 | ···················· | ···················· | ···················· | ···················· | ···················· | ···················· |
| Q | 4014 | hCoV-19/England/MILK-AC7EB6/2020\|EPI_ISL_634350\|20 | ···················· | ···················· | ···················· | ···················· | ···················· | ···················· |
| Q | 4015 | hCoV-19/England/MILK-ACD560/2020\|EPI_ISL_634351\|20 | ···················· | ···················· | ···················· | ···················· | ···················· | ···················· |
| Q | 4016 | hCoV-19/England/MILK-ACD9DD/2020\|EPI_ISL_634352\|20 | ···················· | ···················· | ···················· | ···················· | ···················· | ···················· |
| Q | 4017 | hCoV-19/England/MILK-AC7B55/2020\|EPI_ISL_634354\|20 | ···················· | ···················· | ···················· | ···················· | ···················· | ···················· |
| Q | 4018 | hCoV-19/England/MILK-ACC5E9/2020\|EPI_ISL_634355\|20 | ···················· | ···················· | ···················· | ···················· | ···················· | ···················· |

| | | | | | | | | | |
|---|---|---|---|---|---|---|---|---|---|
| Q | 4019 hCoV-19/England/MILK-ACD463/2020\|EPI_ISL_634356\|20 | ···················· | ···················· | ···················· | ···················· | ······T············· | ···················· | | |
| Q | 4020 hCoV-19/England/MILK-ACC446/2020\|EPI_ISL_634357\|20 | ···················· | ···················· | ···················· | ···················· | ···················· | ···················· | | |
| Q | 4021 hCoV-19/England/MILK-ACD4BE/2020\|EPI_ISL_634358\|20 | ···················· | ···················· | ···················· | ···················· | ···················· | ···················· | | |
| Q | 4022 hCoV-19/England/MILK-ACD481/2020\|EPI_ISL_634359\|20 | ···················· | ···················· | ···················· | ···················· | ···················· | ···················· | | |
| Q | 4023 hCoV-19/USA/CA-ALSR-2634/2020\|EPI_ISL_635600\|2020- | ···················· | ·A·················· | ···················· | ···················· | ···················· | ···················· | | |
| Q | 4024 hCoV-19/USA/CA-ALSR-2635/2020\|EPI_ISL_635601\|2020- | ···················· | ···················· | ···················· | ···················· | ···················· | ···················· | | |
| Q | 4025 hCoV-19/USA/CA-ALSR-2636/2020\|EPI_ISL_635602\|2020- | ···················· | ·A·················· | ···················· | ···················· | ···················· | ···················· | | |
| Q | 4026 hCoV-19/USA/CA-ALSR-2637/2020\|EPI_ISL_635603\|2020- | ···················· | ···················· | ···················· | ···················· | ···················· | ···················· | | |
| Q | 4027 hCoV-19/USA/CA-ALSR-2730/2020\|EPI_ISL_635648\|2020- | ···················· | ···················· | ···················· | ···················· | ···················· | ···················· | | |
| Q | 4028 hCoV-19/USA/CA-ALSR-2733/2020\|EPI_ISL_635649\|2020- | ···················· | ···················· | ···················· | ···················· | ···················· | ···················· | | |
| Q | 4029 hCoV-19/USA/CA-ALSR-2737/2020\|EPI_ISL_635651\|2020- | ···················· | ·A·················· | ···················· | ···················· | ···················· | ···················· | | |
| Q | 4030 hCoV-19/USA/CA-ALSR-2739/2020\|EPI_ISL_635652\|2020- | ···················· | ·A·················· | ···················· | ···················· | ···················· | ···················· | | |
| Q | 4031 hCoV-19/USA/CA-ALSR-2740/2020\|EPI_ISL_635653\|2020- | ···················· | ·A·················· | ···················· | ···················· | ···················· | ···················· | | |
| Q | 4032 hCoV-19/USA/CA-ALSR-2741/2020\|EPI_ISL_635654\|2020- | ···················· | ·A·················· | ···················· | ···················· | ···················· | ···················· | | |
| Q | 4033 hCoV-19/USA/CA-ALSR-2744/2020\|EPI_ISL_635655\|2020- | ···················· | ·A·················· | ···················· | ···················· | ···················· | ···················· | | |
| Q | 4034 hCoV-19/USA/CA-ALSR-2747/2020\|EPI_ISL_635657\|2020- | ···················· | ·A·················· | ···················· | ···················· | ···················· | ···················· | | |
| Q | 4035 hCoV-19/USA/CA-ALSR-2748/2020\|EPI_ISL_635658\|2020- | ···················· | ···················· | ···················· | ···················· | ···················· | ···················· | | |
| Q | 4036 hCoV-19/USA/CA-ALSR-2750/2020\|EPI_ISL_635660\|2020- | ···················· | ·A·················· | ···················· | ···················· | ···················· | ···················· | | |
| Q | 4037 hCoV-19/USA/CA-ALSR-2752/2020\|EPI_ISL_635661\|2020- | ···················· | ·A·················· | ···················· | ···················· | ···················· | ···················· | | |
| Q | 4038 hCoV-19/USA/CA-ALSR-2755/2020\|EPI_ISL_635663\|2020- | ···················· | ·A·················· | ···················· | ···················· | ···················· | ···················· | | |
| Q | 4039 hCoV-19/USA/CA-ALSR-2761/2020\|EPI_ISL_635664\|2020- | ···················· | ···················· | ···················· | ···················· | ···················· | ···················· | | |
| Q | 4040 hCoV-19/USA/CA-ALSR-2767/2020\|EPI_ISL_635668\|2020- | ···················· | ·A·················· | ···················· | ···················· | ···················· | ···················· | | |
| Q | 4041 hCoV-19/USA/CA-ALSR-2768/2020\|EPI_ISL_635669\|2020- | ···················· | ·A·················· | ···················· | ···················· | ···················· | ···················· | | |
| Q | 4042 hCoV-19/USA/CA-ALSR-2769/2020\|EPI_ISL_635670\|2020- | ···················· | ···················· | ···················· | ···················· | ···················· | ···················· | | |
| Q | 4043 hCoV-19/USA/CA-ALSR-2770/2020\|EPI_ISL_635671\|2020- | ···················· | ···················· | ···················· | ···················· | ···················· | ···················· | | |
| Q | 4044 hCoV-19/USA/CA-ALSR-2772/2020\|EPI_ISL_635672\|2020- | ···················· | ·A·················· | ···················· | ···················· | ···················· | ···················· | | |
| Q | 4045 hCoV-19/USA/CA-ALSR-2777/2020\|EPI_ISL_635675\|2020- | ···················· | ·A·················· | ···················· | ···················· | ···················· | ···················· | | |
| Q | 4046 hCoV-19/USA/CA-ALSR-2781/2020\|EPI_ISL_635676\|2020- | ···················· | ···················· | ···················· | ···················· | ···················· | ···················· | | |
| Q | 4047 hCoV-19/USA/CA-ALSR-2784/2020\|EPI_ISL_635677\|2020- | ···················· | ·A·················· | ···················· | ···················· | ···················· | ···················· | | |
| Q | 4048 hCoV-19/USA/CA-ALSR-2786/2020\|EPI_ISL_635678\|2020- | ···················· | ·A·················· | ···················· | ···················· | ···················· | ···················· | | |
| Q | 4049 hCoV-19/USA/CA-ALSR-2792/2020\|EPI_ISL_635680\|2020- | ···················· | ·A·················· | ···················· | ···················· | ···················· | ···················· | | |
| Q | 4050 hCoV-19/USA/CA-ALSR-2794/2020\|EPI_ISL_635681\|2020- | ···················· | ·A·················· | ···················· | ···················· | ···················· | ···················· | | |
| Q | 4051 hCoV-19/USA/CA-ALSR-2800/2020\|EPI_ISL_635683\|2020- | ···················· | ···················· | ···················· | ···················· | ···················· | ···················· | | |
| Q | 4052 hCoV-19/USA/CA-ALSR-2801/2020\|EPI_ISL_635684\|2020- | ···················· | ·A·················· | ···················· | ···················· | ···················· | ···················· | | |
| Q | 4053 hCoV-19/USA/CA-ALSR-2802/2020\|EPI_ISL_635685\|2020- | ···················· | ·A·················· | ···················· | ···················· | ···················· | ···················· | | |
| Q | 4054 hCoV-19/USA/CA-ALSR-2803/2020\|EPI_ISL_635686\|2020- | ···················· | ···················· | ···················· | ···················· | ···················· | ···················· | | |
| Q | 4055 hCoV-19/USA/CA-ALSR-2806/2020\|EPI_ISL_635688\|2020- | ···················· | ·A·················· | ···················· | ···················· | ···················· | ···················· | | |
| Q | 4056 hCoV-19/USA/CA-ALSR-2808/2020\|EPI_ISL_635689\|2020- | ···················· | ·A·················· | ···················· | ···················· | ···················· | ···················· | | |
| Q | 4057 hCoV-19/Lithuania/MR-LUHS-Eilnr470/2020\|EPI_ISL_63 | ···················· | ···················· | ···················· | ···················· | ···················· | ···················· | | |
| Q | 4058 hCoV-19/Lithuania/MR-LUHS-Eilnr471/2020\|EPI_ISL_63 | ···················· | ···················· | ···················· | ···················· | ···················· | ···················· | | |
| Q | 4059 hCoV-19/Lithuania/MR-LUHS-Eilnr474/2020\|EPI_ISL_63 | ···················· | ···················· | ···················· | ···················· | ···················· | ···················· | | |
| Q | 4060 hCoV-19/Lithuania/MR-LUHS-Eilnr475/2020\|EPI_ISL_63 | ···················· | ···················· | ···················· | ···················· | ···················· | ···················· | | |
| Q | 4061 hCoV-19/Lithuania/MR-LUHS-Eilnr476/2020\|EPI_ISL_63 | ···················· | ···················· | ···················· | ···················· | ···················· | ···················· | | |
| Q | 4062 hCoV-19/Lithuania/MR-LUHS-Eilnr477/2020\|EPI_ISL_63 | ···················· | ···················· | ···················· | ···················· | ···················· | ···················· | | |
| Q | 4063 hCoV-19/Lithuania/MR-LUHS-Eilnr479/2020\|EPI_ISL_63 | ···················· | ···················· | ···················· | ···················· | ···················· | ···················· | | |
| Q | 4064 hCoV-19/Canada/ON-PHL-20-01754/2020\|EPI_ISL_636907 | ···················· | ···················· | ···················· | ···················· | ···················· | ···················· | | |
| Q | 4065 hCoV-19/Canada/ON-PHL-20-01755/2020\|EPI_ISL_636908 | ···················· | ···················· | ···················· | ···················· | N··················· | ···················· | | |
| Q | 4068 hCoV-19/Canada/ON-PHL-20-01758/2020\|EPI_ISL_636911 | ···················· | ······A············· | ···················· | ···················· | ···················· | ···················· | | |
| Q | 4069 hCoV-19/Canada/ON-PHL-20-01759/2020\|EPI_ISL_636912 | ···················· | ···················· | ···················· | ···················· | ···················· | ···················· | | |
| Q | 4070 hCoV-19/Canada/ON-PHL-20-01760/2020\|EPI_ISL_636913 | ···················· | ······A············· | ···················· | ···················· | NNN················· | ···················· | | 1 |
| Q | 4071 hCoV-19/Canada/ON-PHL-20-01761/2020\|EPI_ISL_636914 | ···················· | ·A·················· | ···················· | ···················· | ···················· | ···················· | | |
| Q | 4072 hCoV-19/Canada/ON-PHL-20-01762/2020\|EPI_ISL_636915 | ···················· | ···················· | ···················· | ···················· | ··················N· | ···················· | 1 | |
| Q | 4073 hCoV-19/Canada/ON-PHL-20-01763/2020\|EPI_ISL_636916 | ···················· | ···················· | ···················· | ···················· | ···················· | ···················· | | |
| Q | 4074 hCoV-19/Canada/ON-PHL-20-01764/2020\|EPI_ISL_636917 | ···················· | ···················· | ···················· | ···················· | ···················· | ···················· | | |
| Q | 4075 hCoV-19/Canada/ON-PHL-20-01765/2020\|EPI_ISL_636918 | ···················· | ···················· | ···················· | ···················· | ···················· | ···················· | | |
| Q | 4077 hCoV-19/Canada/ON-PHL-20-01767/2020\|EPI_ISL_636920 | ···················· | ···················· | ···················· | ···················· | ···················· | ···················· | | |
| Q | 4079 hCoV-19/Canada/ON-PHL-20-01769/2020\|EPI_ISL_636922 | ···················· | ······A············· | ···················· | ···················· | ···················· | ···················· | | |
| Q | 4080 hCoV-19/Canada/ON-PHL-20-01770/2020\|EPI_ISL_636923 | ···················· | ···················· | ···················· | ···················· | ·NN················· | ···················· | | |
| Q | 4081 hCoV-19/Canada/ON-PHL-20-01771/2020\|EPI_ISL_636924 | ···················· | ······A············· | ···················· | ···················· | ···················· | ···················· | | |
| Q | 4084 hCoV-19/Canada/ON-PHL-20-01776/2020\|EPI_ISL_636928 | ···················· | ···················· | ···················· | ···················· | N··················· | ···················· | | |
| Q | 4086 hCoV-19/Canada/ON-PHL-20-01778/2020\|EPI_ISL_636930 | ···················· | ···················· | ···················· | ···················· | NNN················· | ···················· | | 1 |
| Q | 4087 hCoV-19/Canada/ON-PHL-20-01780/2020\|EPI_ISL_636931 | ···················· | ···················· | ···················· | ···················· | ···················· | ···················· | | |
| Q | 4090 hCoV-19/Canada/ON-PHL-20-01783/2020\|EPI_ISL_636934 | ···················· | ···················· | ···················· | ···················· | ···················· | ···················· | | |
| Q | 4091 hCoV-19/Mozambique/KRISP-MZ158905/2020\|EPI_ISL_636 | ···················· | ···················· | ···················· | ···················· | ···················· | ···················· | | |
| Q | 4092 hCoV-19/DRC/D07-1216/2020\|EPI_ISL_636982\|2020-05-2 | ···················· | ···················· | ···················· | ···················· | ···················· | ···················· | | |
| Q | 4093 hCoV-19/DRC/D09-0676/2020\|EPI_ISL_636983\|2020-05-0 | ···················· | ···················· | ···················· | ···················· | ···················· | ···················· | | |
| Q | 4094 hCoV-19/DRC/D09-0678/2020\|EPI_ISL_636984\|2020-05-1 | ···················· | ···················· | ···················· | ···················· | ···················· | ···················· | | |
| Q | 4095 hCoV-19/DRC/D09-0684/2020\|EPI_ISL_636985\|2020-05-1 | ···················· | ···················· | ···················· | ···················· | ···················· | ···················· | | |
| Q | 4096 hCoV-19/DRC/D09-0687/2020\|EPI_ISL_636986\|2020-05-2 | ···················· | ··············G····· | ···················· | ···················· | ···················· | ···················· | | |
| Q | 4097 hCoV-19/Wales/QEUH-A49960/2020\|EPI_ISL_628440\|2020 | ···················· | ···················· | ···················· | ···················· | ···················· | ···················· | | |
| Q | 4098 hCoV-19/Scotland/QEUH-A4997F/2020\|EPI_ISL_628441\|2 | ···················· | ···················· | ···················· | ···················· | ···················· | ···················· | | |
| Q | 4099 hCoV-19/Scotland/QEUH-A4998E/2020\|EPI_ISL_628442\|2 | ···················· | ···················· | ···················· | ···················· | ···················· | ···················· | | |
| Q | 4100 hCoV-19/Scotland/QEUH-A499BB/2020\|EPI_ISL_628443\|2 | ···················· | ···················· | ···················· | ···················· | ···················· | ···················· | | |
| Q | 4101 hCoV-19/Scotland/QEUH-A499F7/2020\|EPI_ISL_628444\|2 | ···················· | ···················· | ···················· | ···················· | ···················· | ···················· | | |
| Q | 4102 hCoV-19/England/QEUH-A49A12/2020\|EPI_ISL_628445\|20 | ···················· | ···················· | ···················· | ···················· | ···················· | ···················· | | |
| Q | 4103 hCoV-19/England/QEUH-A49A21/2020\|EPI_ISL_628446\|20 | ···················· | ···················· | ···················· | ···················· | ···················· | ···················· | | |
| Q | 4104 hCoV-19/England/QEUH-A49A4F/2020\|EPI_ISL_628447\|20 | ···················· | ···················· | ···················· | ···················· | ···················· | ···················· | | |
| Q | 4105 hCoV-19/England/QEUH-A49A7C/2020\|EPI_ISL_628448\|20 | ···················· | ···················· | ···················· | ···················· | ···················· | ···················· | | |
| Q | 4106 hCoV-19/England/QEUH-A49A8B/2020\|EPI_ISL_628449\|20 | ···················· | ···················· | ···················· | ···················· | ···················· | ···················· | | |
| Q | 4107 hCoV-19/England/QEUH-A49A9A/2020\|EPI_ISL_628450\|20 | ···················· | ···················· | ···················· | ···················· | ···················· | ···················· | | |

| | | | | | | | |
|---|---|---|---|---|---|---|---|
| Q | 4108 hCoV-19/England/QEUH-A49AA9/2020\|EPI_ISL_628451\|20 | ···················· | ···················· | ···················· | ···················· | ···················· | ···················· |
| Q | 4109 hCoV-19/England/QEUH-A49AC7/2020\|EPI_ISL_628452\|20 | ···················· | ···················· | ···················· | ···················· | ···················· | ···················· |
| Q | 4110 hCoV-19/England/QEUH-A49AD6/2020\|EPI_ISL_628453\|20 | ···················· | ···················· | ···················· | ···················· | ···················· | ···················· |
| Q | 4111 hCoV-19/England/QEUH-A49B1F/2020\|EPI_ISL_628454\|20 | ···················· | ···················· | ···················· | ···················· | ···················· | ···················· |
| Q | 4112 hCoV-19/Scotland/QEUH-A49B6A/2020\|EPI_ISL_628455\|2 | ···················· | ···················· | ···················· | ···················· | ···················· | ···················· |
| Q | 4113 hCoV-19/Scotland/QEUH-A4AE51/2020\|EPI_ISL_628456\|2 | ···················· | ···················· | ···················· | ···················· | ···················· | ···················· |
| Q | 4114 hCoV-19/Scotland/QEUH-A4AE7F/2020\|EPI_ISL_628457\|2 | ···················· | ···················· | ···················· | ···················· | ···················· | ···················· |
| Q | 4115 hCoV-19/England/QEUH-A4AE9D/2020\|EPI_ISL_628458\|20 | ···················· | ···················· | ···················· | ···················· | ···················· | ···················· |
| Q | 4116 hCoV-19/England/QEUH-A4AECA/2020\|EPI_ISL_628459\|20 | ···················· | ···················· | ···················· | ···················· | ···················· | ···················· |
| Q | 4117 hCoV-19/England/MILK-AC8204/2020\|EPI_ISL_629700\|20 | ···················· | ···················· | ···················· | ···················· | ···················· | ···················· |
| Q | 4118 hCoV-19/England/MILK-ACEAE8/2020\|EPI_ISL_629704\|20 | ···················· | ···················· | ···················· | ···················· | ···················· | ···················· |
| Q | 4119 hCoV-19/England/MILK-ACF20D/2020\|EPI_ISL_629705\|20 | ···················· | ···················· | ···················· | ···················· | ···················· | ···················· |
| Q | 4120 hCoV-19/England/MILK-AC8F3A/2020\|EPI_ISL_629707\|20 | ···················· | ···················· | ···················· | ···················· | ···················· | ···················· |
| Q | 4121 hCoV-19/England/MILK-ACEA24/2020\|EPI_ISL_629708\|20 | ···················· | ·A·················· | ···················· | ···················· | ···················· | ···················· |
| Q | 4122 hCoV-19/England/MILK-ACF3DD/2020\|EPI_ISL_629709\|20 | ···················· | ···················· | ···················· | ···················· | ···················· | ···················· |
| Q | 4123 hCoV-19/England/MILK-AC8F94/2020\|EPI_ISL_629712\|20 | ···················· | ···················· | ···················· | ···················· | ·····T·············· | ···················· |
| Q | 4124 hCoV-19/England/MILK-ACF2B2/2020\|EPI_ISL_629713\|20 | ···················· | ···················· | ···················· | ···················· | ···················· | ···················· |
| Q | 4125 hCoV-19/England/MILK-AC8F85/2020\|EPI_ISL_629714\|20 | ···················· | ···················· | ···················· | ···················· | ···················· | ···················· |
| Q | 4126 hCoV-19/England/MILK-ACFA6F/2020\|EPI_ISL_629715\|20 | ···················· | ···················· | ···················· | ···················· | ···················· | ···················· |
| Q | 4127 hCoV-19/England/MILK-ACF57D/2020\|EPI_ISL_629716\|20 | ···················· | ···················· | ···················· | ···················· | ·····T·············· | ···················· |
| Q | 4128 hCoV-19/England/MILK-ACEACA/2020\|EPI_ISL_629717\|20 | ···················· | ···················· | ···················· | ···················· | ···················· | ···················· |
| Q | 4129 hCoV-19/England/MILK-AC91E8/2020\|EPI_ISL_629718\|20 | ···················· | ···················· | ···················· | ···················· | ···················· | ···················· |
| Q | 4130 hCoV-19/England/MILK-AC825F/2020\|EPI_ISL_629719\|20 | ···················· | ···················· | ···················· | ···················· | ···················· | ···················· |
| Q | 4131 hCoV-19/England/MILK-ACEFF8/2020\|EPI_ISL_629721\|20 | ···················· | ···················· | ···················· | ···················· | ···················· | ···················· |
| Q | 4132 hCoV-19/England/MILK-ACE2B3/2020\|EPI_ISL_629722\|20 | ···················· | ···················· | ···················· | ···················· | ···················· | ···················· |
| Q | 4133 hCoV-19/England/MILK-ACE620/2020\|EPI_ISL_629724\|20 | ···················· | ···················· | ···················· | ···················· | ···················· | ···················· |
| Q | 4134 hCoV-19/England/MILK-ACFA41/2020\|EPI_ISL_629726\|20 | ···················· | ···················· | ···················· | ···················· | ···················· | ···················· |
| Q | 4135 hCoV-19/England/QEUH-AD2F4C/2020\|EPI_ISL_629770\|20 | ···················· | ···················· | ···················· | ···················· | ···················· | ···················· |
| Q | 4136 hCoV-19/England/QEUH-AD1EE6/2020\|EPI_ISL_629771\|20 | ···················· | ···················· | ···················· | ···················· | ···················· | ···················· |
| Q | 4137 hCoV-19/England/QEUH-AD24B6/2020\|EPI_ISL_629772\|20 | ···················· | ···················· | ···················· | ···················· | ···················· | ···················· |
| Q | 4138 hCoV-19/England/QEUH-AD2E03/2020\|EPI_ISL_629773\|20 | ···················· | ···················· | ···················· | ···················· | ···················· | ···················· |
| Q | 4139 hCoV-19/England/QEUH-AD0430/2020\|EPI_ISL_629774\|20 | ···················· | ···················· | ···················· | ···················· | ···················· | ···················· |
| Q | 4140 hCoV-19/England/QEUH-AD1E5F/2020\|EPI_ISL_629775\|20 | ···················· | ···················· | ···················· | ···················· | ···················· | ···················· |
| Q | 4141 hCoV-19/England/QEUH-AD21FB/2020\|EPI_ISL_629776\|20 | ···················· | ···················· | ···················· | ···················· | ···················· | ···················· |
| Q | 4142 hCoV-19/England/QEUH-AD03CA/2020\|EPI_ISL_629777\|20 | ···················· | ·A·················· | ···················· | ···················· | ···················· | ··A················· |
| Q | 4143 hCoV-19/England/QEUH-AD3E02/2020\|EPI_ISL_629778\|20 | ···················· | ···················· | ···················· | ···················· | ···················· | ···················· |
| Q | 4144 hCoV-19/England/QEUH-AD1DF8/2020\|EPI_ISL_629779\|20 | ···················· | ···················· | ···················· | ···················· | ···················· | ···················· |
| Q | 4145 hCoV-19/England/QEUH-AD253B/2020\|EPI_ISL_629780\|20 | ···················· | ···················· | ···················· | ···················· | ···················· | ···················· |
| Q | 4146 hCoV-19/England/QEUH-AD0676/2020\|EPI_ISL_629781\|20 | ···················· | ·A·················· | ···················· | ···················· | ···················· | ···················· |
| Q | 4147 hCoV-19/England/QEUH-AD62D6/2020\|EPI_ISL_629782\|20 | ···················· | ···················· | ···················· | ···················· | ···················· | ···················· |
| Q | 4148 hCoV-19/England/QEUH-AD2771/2020\|EPI_ISL_629785\|20 | ···················· | ···················· | ···················· | ···················· | ···················· | ···················· |
| Q | 4149 hCoV-19/England/QEUH-AD2F88/2020\|EPI_ISL_629786\|20 | ···················· | ···················· | ···················· | ···················· | ···················· | ···················· |
| Q | 4150 hCoV-19/England/QEUH-AD1D43/2020\|EPI_ISL_629787\|20 | ···················· | ···················· | ···················· | ···················· | ···················· | ···················· |
| Q | 4151 hCoV-19/England/QEUH-AD2F79/2020\|EPI_ISL_629789\|20 | ···················· | ···················· | ···················· | ···················· | ···················· | ···················· |
| Q | 4152 hCoV-19/England/ALDP-A670B2/2020\|EPI_ISL_630700\|20 | ···················· | ···················· | ···················· | ···················· | ···················· | ···················· |
| Q | 4153 hCoV-19/England/ALDP-A672BC/2020\|EPI_ISL_630701\|20 | ···················· | ·A·················· | ···················· | ···················· | ···················· | ··A················· |
| Q | 4154 hCoV-19/England/QEUH-AAF2B8/2020\|EPI_ISL_630702\|20 | ···················· | ···················· | ···················· | ···················· | ···················· | ···················· |
| Q | 4155 hCoV-19/England/QEUH-AAE77E/2020\|EPI_ISL_630703\|20 | ···················· | ···················· | ···················· | ···················· | ···················· | ···················· |
| Q | 4156 hCoV-19/England/QEUH-AAF090/2020\|EPI_ISL_630704\|20 | ···················· | ···················· | ···················· | ···················· | ···················· | ···················· |
| Q | 4157 hCoV-19/England/QEUH-AAF300/2020\|EPI_ISL_630705\|20 | ···················· | ···················· | ···················· | ···················· | ···················· | ···················· |
| Q | 4158 hCoV-19/England/ALDP-AA4ECE/2020\|EPI_ISL_630707\|20 | ···················· | ···················· | ···················· | ···················· | ···················· | ···················· |
| Q | 4159 hCoV-19/England/ALDP-AA511F/2020\|EPI_ISL_630708\|20 | ···················· | ···················· | ···················· | ···················· | ···················· | ···················· |
| Q | 4160 hCoV-19/England/ALDP-AA4EFB/2020\|EPI_ISL_630709\|20 | ···················· | ···················· | ···················· | ···················· | ···················· | ···················· |
| Q | 4161 hCoV-19/England/ALDP-AA507C/2020\|EPI_ISL_630753\|20 | ···················· | ···················· | ···················· | ···················· | ···················· | ···················· |
| Q | 4162 hCoV-19/England/ALDP-AA50D6/2020\|EPI_ISL_630754\|20 | ···················· | ···················· | ···················· | ···················· | ···················· | ···················· |
| Q | 4163 hCoV-19/England/ALDP-AA4CA6/2020\|EPI_ISL_630755\|20 | ···················· | ···················· | ···················· | ···················· | ···················· | ···················· |
| Q | 4164 hCoV-19/England/ALDP-AA291A/2020\|EPI_ISL_630756\|20 | ···················· | ···················· | ···················· | ···················· | ···················· | ···················· |
| Q | 4165 hCoV-19/England/ALDP-AA25BC/2020\|EPI_ISL_630757\|20 | ···················· | ···················· | ···················· | ···················· | ···················· | ···················· |
| Q | 4166 hCoV-19/England/ALDP-AA28E0/2020\|EPI_ISL_630758\|20 | ···················· | ···················· | ···················· | ···················· | ···················· | ···················· |
| Q | 4167 hCoV-19/England/ALDP-AA514C/2020\|EPI_ISL_630759\|20 | ···················· | ···················· | ···················· | ···················· | ···················· | ···················· |
| Q | 4168 hCoV-19/England/ALDP-AA31C6/2020\|EPI_ISL_630760\|20 | ···················· | ···················· | ···················· | ···················· | ···················· | ···················· |
| Q | 4169 hCoV-19/England/QEUH-AAE073/2020\|EPI_ISL_630761\|20 | ···················· | ···················· | ···················· | ···················· | ···················· | ···················· |
| Q | 4170 hCoV-19/England/ALDP-AA31E4/2020\|EPI_ISL_630763\|20 | ···················· | ···················· | ···················· | ···················· | ···················· | ···················· |
| Q | 4171 hCoV-19/England/QEUH-AAE240/2020\|EPI_ISL_630764\|20 | ···················· | ···················· | ···················· | ···················· | ···T················ | ···················· |
| Q | 4172 hCoV-19/England/ALDP-AA4ACA/2020\|EPI_ISL_630766\|20 | ···················· | ···················· | ···················· | ···················· | ···················· | ···················· |
| Q | 4173 hCoV-19/England/ALDP-AA4EDD/2020\|EPI_ISL_630767\|20 | ···················· | ···················· | ···················· | ···················· | ···················· | ···················· |
| Q | 4174 hCoV-19/England/ALDP-AA25DA/2020\|EPI_ISL_630768\|20 | ···················· | ···················· | ···················· | ···················· | ···················· | ···················· |
| Q | 4175 hCoV-19/England/ALDP-AA09B2/2020\|EPI_ISL_630769\|20 | ···················· | ···················· | ···················· | ···················· | ···················· | ···················· |
| Q | 4176 hCoV-19/England/ALDP-AA2A44/2020\|EPI_ISL_630772\|20 | ···················· | ···················· | ···················· | ···················· | ···················· | ···················· |
| Q | 4177 hCoV-19/England/ALDP-AA26B9/2020\|EPI_ISL_630773\|20 | ···················· | ···················· | ···················· | ···················· | ···················· | ···················· |
| Q | 4178 hCoV-19/England/ALDP-AA269B/2020\|EPI_ISL_630774\|20 | ···················· | ···················· | ···················· | ···················· | ···················· | ···················· |
| Q | 4179 hCoV-19/USA/NY-NYCPHL-000981/2020\|EPI_ISL_633008\|2 | ···················· | ···················· | ···················· | ···················· | ···················· | ···················· |
| Q | 4180 hCoV-19/USA/NY-NYCPHL-000982/2020\|EPI_ISL_633009\|2 | ···················· | ·A·················· | ···················· | ···················· | ···················· | ···················· |
| Q | 4181 hCoV-19/USA/NY-NYCPHL-000985/2020\|EPI_ISL_633011\|2 | ···················· | ···················· | ···················· | ···················· | ···················· | ···················· |
| Q | 4182 hCoV-19/USA/NY-NYCPHL-000986/2020\|EPI_ISL_633012\|2 | ···················· | ···················· | ···················· | ···················· | ···················· | ···················· |
| Q | 4183 hCoV-19/England/QEUH-AD525F/2020\|EPI_ISL_634240\|20 | ···················· | ···················· | ···················· | ···················· | ···················· | ···················· |
| Q | 4184 hCoV-19/England/QEUH-AD34F1/2020\|EPI_ISL_634241\|20 | ···················· | ·A·················· | ···················· | ···················· | ···················· | ··A················· |
| Q | 4185 hCoV-19/England/QEUH-AD2CAF/2020\|EPI_ISL_634243\|20 | ···················· | ···················· | ···················· | ···················· | ···················· | ···················· |
| Q | 4186 hCoV-19/England/QEUH-AD0D35/2020\|EPI_ISL_634244\|20 | ···················· | ···················· | ···················· | ···················· | ···················· | ···················· |
| Q | 4187 hCoV-19/England/QEUH-AD30FD/2020\|EPI_ISL_634245\|20 | ···················· | ···················· | ···················· | ···················· | ···················· | ···················· |

Q 4188 hCoV-19/England/QEUH-AD2CEB/2020|EPI_ISL_634246|20 ···················· ···················· ····················· ···················· ···················· ····················
Q 4189 hCoV-19/England/QEUH-AD51DA/2020|EPI_ISL_634247|20 ···················· ···················· ····················· ···················· ···················· ····················
Q 4190 hCoV-19/England/QEUH-AD1FB6/2020|EPI_ISL_634248|20 ···················· ···················· ····················· ···················· ···················· ····················
Q 4191 hCoV-19/England/QEUH-AD037F/2020|EPI_ISL_634249|20 ···················· ···················· ····················· ···················· ···················· ····················
Q 4192 hCoV-19/England/QEUH-AD01B1/2020|EPI_ISL_634250|20 ···················· ···················· ····················· ···················· ···················· ····················
Q 4193 hCoV-19/England/QEUH-AD5055/2020|EPI_ISL_634252|20 ···················· ···················· ····················· ···················· ···················· ····················
Q 4194 hCoV-19/England/MILK-ACC947/2020|EPI_ISL_634256|20 ···················· ···················· ····················· ···················· ···················· ····················
Q 4195 hCoV-19/England/MILK-ACC6F5/2020|EPI_ISL_634258|20 ···················· ···················· ····················· ···················· ···················· ····················
Q 4196 hCoV-19/England/MILK-ACC482/2020|EPI_ISL_634259|20 ···················· ···················· ····················· ···················· ···················· ····················
Q 4197 hCoV-19/England/MILK-AC8134/2020|EPI_ISL_634261|20 ···················· ···················· ····················· ···················· ···················· ····················
Q 4198 hCoV-19/England/MILK-ACC5F8/2020|EPI_ISL_634262|20 ···················· ···················· ····················· ···················· ···················· ····················
Q 4199 hCoV-19/England/MILK-ACC6AA/2020|EPI_ISL_634263|20 ···················· ···················· ····················· ···················· ···················· ····················
Q 4200 hCoV-19/England/MILK-ACD59D/2020|EPI_ISL_634264|20 ···················· ···················· ····················· ···················· ···················· ····················
Q 4201 hCoV-19/England/MILK-AC7A1C/2020|EPI_ISL_634265|20 ···················· ···················· ····················· ···················· ···················· ····················
Q 4202 hCoV-19/England/MILK-ACC7F2/2020|EPI_ISL_634267|20 ···················· ···················· ····················· ···················· ···················· ····················
Q 4203 hCoV-19/England/MILK-ACD490/2020|EPI_ISL_634268|20 ···················· ···················· ····················· ···················· ···················· ····················
Q 4204 hCoV-19/England/MILK-ACC570/2020|EPI_ISL_634269|20 ···················· ···················· ····················· ···················· ···················· ····················
Q 4205 hCoV-19/England/MILK-ACC5DA/2020|EPI_ISL_634270|20 ···················· ·A·················· ····················· ···················· ···················· ··A·················
Q 4206 hCoV-19/England/MILK-ACC84A/2020|EPI_ISL_634271|20 ···················· ···················· ····················· ···················· ···················· ····················
Q 4207 hCoV-19/England/MILK-ACCEC0/2020|EPI_ISL_634272|20 ···················· ···················· ····················· ···················· ···················· ····················
Q 4208 hCoV-19/England/MILK-ACD4CD/2020|EPI_ISL_634273|20 ···················· ···················· ····················· ···················· ···················· ····················
Q 4209 hCoV-19/England/MILK-ACC6C8/2020|EPI_ISL_634274|20 ···················· ···················· ····················· ···················· ···················· ····················
Q 4210 hCoV-19/England/MILK-AC7C16/2020|EPI_ISL_634275|20 ···················· ···················· ····················· ···················· ···················· ····················
Q 4211 hCoV-19/England/MILK-AC7E4D/2020|EPI_ISL_634276|20 ···················· ···················· ····················· ···················· ···················· ····················
Q 4212 hCoV-19/England/MILK-AC7C07/2020|EPI_ISL_634277|20 ···················· ···················· ····················· ···················· ···················· ····················
Q 4213 hCoV-19/England/MILK-AC79D3/2020|EPI_ISL_634278|20 ···················· ·A·················· ····················· ···················· ···················· ··A·················
Q 4214 hCoV-19/England/MILK-ACD515/2020|EPI_ISL_634280|20 ···················· ···················· ····················· ···················· ···················· ····················
Q 4215 hCoV-19/England/MILK-ACC80E/2020|EPI_ISL_634281|20 ···················· ···················· ····················· ···················· ···················· ····················
Q 4216 hCoV-19/England/MILK-AC7BBF/2020|EPI_ISL_634282|20 ···················· ···················· ····················· ···················· ···················· ····················
Q 4217 hCoV-19/England/MILK-ACD66D/2020|EPI_ISL_634283|20 ···················· ·A·················· ····················· ···················· ···················· ··A·················
Q 4218 hCoV-19/England/MILK-AC80BF/2020|EPI_ISL_634284|20 ···················· ···················· ····················· ···················· ···················· ····················
Q 4219 hCoV-19/England/MILK-ACC6D7/2020|EPI_ISL_634285|20 ···················· ···················· ····················· ···················· ···················· ····················
Q 4220 hCoV-19/England/MILK-AC789A/2020|EPI_ISL_634286|20 ···················· ···················· ····················· ···················· ···················· ····················
Q 4221 hCoV-19/England/MILK-ACD69A/2020|EPI_ISL_634287|20 ···················· ·A·················· ····················· ···················· ···················· ··A·················
Q 4222 hCoV-19/England/MILK-ACCBBA/2020|EPI_ISL_634288|20 ···················· ···················· ····················· ···················· ···················· ····················
Q 4223 hCoV-19/England/MILK-ACCEB1/2020|EPI_ISL_634289|20 ···················· ·A·················· ····················· ···················· ···················· ··A·················
Q 4224 hCoV-19/England/MILK-ACD1F3/2020|EPI_ISL_634290|20 ···················· ···················· ····················· ···················· ···················· ····················
Q 4225 hCoV-19/England/MILK-ACC455/2020|EPI_ISL_634291|20 ···················· ···················· ····················· ···················· ···················· ····················
Q 4226 hCoV-19/England/MILK-ACD68B/2020|EPI_ISL_634292|20 ···················· ···················· ····················· ···················· ···················· ····················
Q 4227 hCoV-19/England/MILK-ACCAAE/2020|EPI_ISL_634293|20 ···················· ···················· ····················· ···················· ···················· ····················
Q 4228 hCoV-19/England/MILK-ACD427/2020|EPI_ISL_634294|20 ···················· ···················· ····················· ···················· ···················· ····················
Q 4229 hCoV-19/England/MILK-ACD269/2020|EPI_ISL_634295|20 ···················· ···················· ····················· ···················· ···················· ····················
Q 4230 hCoV-19/England/MILK-ACD1E4/2020|EPI_ISL_634296|20 ···················· ···················· ····················· ···················· ···················· ····················
Q 4231 hCoV-19/England/MILK-ACC983/2020|EPI_ISL_634297|20 ···················· ···················· ····················· ···················· ···················· ····················
Q 4232 hCoV-19/England/MILK-AC7AD0/2020|EPI_ISL_634298|20 ···················· ···················· ····················· ···················· ···················· ····················
Q 4233 hCoV-19/England/MILK-AC7B64/2020|EPI_ISL_634299|20 ···················· ···················· ····················· ···················· ···················· ····················
Q 4234 hCoV-19/England/MILK-AC78A9/2020|EPI_ISL_634300|20 ···················· ···················· ····················· ···················· ···················· ····················
Q 4235 hCoV-19/England/MILK-ACCBE7/2020|EPI_ISL_634303|20 ···················· ···················· ····················· ···················· ···················· ····················
Q 4236 hCoV-19/England/MILK-AD002D/2020|EPI_ISL_634305|20 ···················· ···················· ····················· ···················· ···················· ····················
Q 4237 hCoV-19/England/MILK-ACC965/2020|EPI_ISL_634306|20 ···················· ···················· ····················· ···················· ···················· ····················
Q 4238 hCoV-19/England/MILK-AC75EE/2020|EPI_ISL_634307|20 ···················· ···················· ····················· ···················· ···················· ····················
Q 4239 hCoV-19/England/MILK-ACD67C/2020|EPI_ISL_634308|20 ···················· ···················· ····················· ···················· ···················· ····················
Q 4240 hCoV-19/England/MILK-AC7609/2020|EPI_ISL_634309|20 ···················· ···················· ····················· ···················· ···················· ····················
Q 4241 hCoV-19/England/MILK-ACC929/2020|EPI_ISL_634310|20 ···················· ···················· ····················· ···················· ···················· ····················
Q 4242 hCoV-19/England/MILK-ACCC02/2020|EPI_ISL_634312|20 ···················· ·A·················· ····················· ···················· ···················· ··A·················
Q 4243 hCoV-19/England/MILK-AC7A49/2020|EPI_ISL_634313|20 ···················· ···················· ····················· ···················· ···················· ····················
Q 4244 hCoV-19/England/MILK-ACD472/2020|EPI_ISL_634314|20 ···················· ···················· ····················· ···················· ···················· ····················
Q 4245 hCoV-19/England/MILK-ACD6B8/2020|EPI_ISL_634315|20 ···················· ···················· ····················· ···················· ···················· ····················
Q 4246 hCoV-19/England/MILK-ACCABD/2020|EPI_ISL_634316|20 ···················· ···················· ····················· ···················· ···················· ····················
Q 4247 hCoV-19/England/MILK-ACD506/2020|EPI_ISL_634342|20 ···················· ·A·················· ····················· ···················· ···················· ··A·················
Q 4248 hCoV-19/England/MILK-ACC868/2020|EPI_ISL_634343|20 ···················· ···················· ····················· ···················· ···················· ····················
Q 4249 hCoV-19/England/MILK-ACCB8D/2020|EPI_ISL_634344|20 ···················· ···················· ····················· ···················· ···················· ····················
Q 4250 hCoV-19/Mexico/BCN-ALSR-2569/2020|EPI_ISL_635570|2 ···················· ·A·················· ····················· ···················· ···················· ····················
Q 4251 hCoV-19/Mexico/BCN-ALSR-2570/2020|EPI_ISL_635571|2 ···················· ·A·················· ····················· ···················· ···················· ····················
Q 4252 hCoV-19/Mexico/BCN-ALSR-2571/2020|EPI_ISL_635572|2 ···················· ·A·················· ····················· ···················· ···················· ····················
Q 4253 hCoV-19/Mexico/BCN-ALSR-2572/2020|EPI_ISL_635573|2 ···················· ···················· ····················· ···················· ···················· ····················
Q 4254 hCoV-19/Mexico/BCN-ALSR-2573/2020|EPI_ISL_635574|2 ···················· ·A·················· ····················· ···················· ···················· ····················
Q 4255 hCoV-19/Mexico/BCN-ALSR-2574/2020|EPI_ISL_635575|2 ···················· ·A·················· ····················· ···················· ···················· ····················
Q 4256 hCoV-19/Mexico/BCN-ALSR-2576/2020|EPI_ISL_635576|2 ···················· ·A·················· ····················· ···················· ···················· ····················
Q 4257 hCoV-19/USA/CA-ALSR-2597/2020|EPI_ISL_635577|2020- ···················· ·A·················· ····················· ···················· ···················· ····················
Q 4258 hCoV-19/USA/CA-ALSR-2598/2020|EPI_ISL_635578|2020- ···················· ···················· ····················· ···················· ···················· ····················
Q 4259 hCoV-19/USA/CA-ALSR-2599/2020|EPI_ISL_635579|2020- ···················· ·A·················· ····················· ···················· ···················· ····················
Q 4260 hCoV-19/USA/CA-ALSR-2600/2020|EPI_ISL_635580|2020- ···················· ·A·················· ····················· ···················· ···················· ····················
Q 4261 hCoV-19/USA/CA-ALSR-2603/2020|EPI_ISL_635582|2020- ···················· ···················· ····················· ···················· ···················· ····················
Q 4262 hCoV-19/USA/CA-ALSR-2605/2020|EPI_ISL_635583|2020- ·······T············ ·A·················· ····················· ···················· ···················· ····················
Q 4263 hCoV-19/USA/CA-ALSR-2607/2020|EPI_ISL_635584|2020- ···················· ·A·················· ····················· ···················· ···················· ····················
Q 4264 hCoV-19/USA/CA-ALSR-2610/2020|EPI_ISL_635585|2020- ·······T············ ·A·················· ····················· ···················· ···················· ····················
Q 4265 hCoV-19/USA/CA-ALSR-2611/2020|EPI_ISL_635586|2020- ···················· ···················· ····················· ···················· ···················· ····················
Q 4266 hCoV-19/USA/CA-ALSR-2614/2020|EPI_ISL_635587|2020- ···················· ·A·················· ····················· ···················· ···················· ····················
Q 4267 hCoV-19/USA/CA-ALSR-2615/2020|EPI_ISL_635588|2020- ···················· ·A·················· ····················· ···················· ···················· ····················

| | | | | | | | | |
|---|---|---|---|---|---|---|---|---|
| Q | 4268 | hCoV-19/USA/CA-ALSR-2616/2020\|EPI_ISL_635589\|2020- | ···················· | ·A·················· | ···················· | ···················· | ···················· | ···················· |
| Q | 4269 | hCoV-19/USA/CA-ALSR-2620/2020\|EPI_ISL_635591\|2020- | ···················· | ···················· | ···················· | ···················· | ···················· | ···················· |
| Q | 4270 | hCoV-19/USA/CA-ALSR-2621/2020\|EPI_ISL_635592\|2020- | ···················· | ···················· | ···················· | ···················· | ···················· | ···················· |
| Q | 4271 | hCoV-19/USA/CA-ALSR-2622/2020\|EPI_ISL_635593\|2020- | ···················· | ···················· | ···················· | ···················· | ···················· | ···················· |
| Q | 4272 | hCoV-19/USA/CA-ALSR-2624/2020\|EPI_ISL_635594\|2020- | ···················· | ···················· | ···················· | ···················· | ···················· | ···················· |
| Q | 4273 | hCoV-19/USA/CA-ALSR-2627/2020\|EPI_ISL_635595\|2020- | ···················· | ·A·················· | ···················· | ···················· | ···················· | ···················· |
| Q | 4274 | hCoV-19/USA/CA-ALSR-2628/2020\|EPI_ISL_635596\|2020- | ···················· | ·A·················· | ···················· | ···················· | ···················· | ···················· |
| Q | 4275 | hCoV-19/USA/CA-ALSR-2629/2020\|EPI_ISL_635597\|2020- | ·······T············ | ·A·················· | ···················· | ···················· | ···················· | ···················· |
| Q | 4276 | hCoV-19/USA/CA-ALSR-2631/2020\|EPI_ISL_635598\|2020- | ···················· | ·A·················· | ···················· | ···················· | ···················· | ···················· |
| Q | 4277 | hCoV-19/USA/CA-ALSR-2638/2020\|EPI_ISL_635604\|2020- | ···················· | ·A·················· | ···················· | ···················· | ···················· | ···················· |
| Q | 4278 | hCoV-19/USA/CA-ALSR-2640/2020\|EPI_ISL_635605\|2020- | ···················· | ·A·················· | ···················· | ···················· | ···················· | ···················· |
| Q | 4279 | hCoV-19/USA/CA-ALSR-2641/2020\|EPI_ISL_635606\|2020- | ···················· | ···················· | ···················· | ···················· | ···················· | ···················· |
| Q | 4280 | hCoV-19/USA/CA-ALSR-2642/2020\|EPI_ISL_635607\|2020- | ···················· | ·A·················· | ···················· | ···················· | ···················· | ···················· |
| Q | 4281 | hCoV-19/USA/CA-ALSR-2643/2020\|EPI_ISL_635608\|2020- | ···················· | ·A·················· | ···················· | ···················· | ···················· | ···················· |
| Q | 4282 | hCoV-19/USA/CA-ALSR-2644/2020\|EPI_ISL_635609\|2020- | ···················· | ·A·················· | ···················· | ···················· | ···················· | ···················· |
| Q | 4283 | hCoV-19/USA/CA-ALSR-2647/2020\|EPI_ISL_635610\|2020- | ···················· | ·A·················· | ···················· | ···················· | ···················· | ···················· |
| Q | 4284 | hCoV-19/USA/CA-ALSR-2652/2020\|EPI_ISL_635611\|2020- | ···················· | ·A·················· | ···················· | ···················· | ···················· | ···················· |
| Q | 4285 | hCoV-19/USA/CA-ALSR-2654/2020\|EPI_ISL_635612\|2020- | ···················· | ·A·················· | ···················· | ···················· | ···················· | ···················· |
| Q | 4286 | hCoV-19/USA/CA-ALSR-2657/2020\|EPI_ISL_635613\|2020- | ···················· | ·A·················· | ···················· | ···················· | ···················· | ···················· |
| Q | 4287 | hCoV-19/USA/CA-ALSR-2659/2020\|EPI_ISL_635614\|2020- | ···················· | ·A·················· | ···················· | ···················· | ···················· | ···················· |
| Q | 4288 | hCoV-19/USA/CA-ALSR-2660/2020\|EPI_ISL_635615\|2020- | ···················· | ···················· | ···················· | ···················· | ···················· | ···················· |
| Q | 4289 | hCoV-19/USA/CA-ALSR-2662/2020\|EPI_ISL_635616\|2020- | ···················· | ·A·················· | ···················· | ···················· | ···················· | ···················· |
| Q | 4290 | hCoV-19/USA/CA-ALSR-2666/2020\|EPI_ISL_635617\|2020- | ···················· | ·A·················· | ···················· | ···················· | ···················· | ···················· |
| Q | 4291 | hCoV-19/USA/CA-ALSR-2667/2020\|EPI_ISL_635618\|2020- | ···················· | ·A·················· | ···················· | ···················· | ···················· | ···················· |
| Q | 4292 | hCoV-19/USA/CA-ALSR-2668/2020\|EPI_ISL_635619\|2020- | ···················· | ···················· | ···················· | ···················· | ···················· | ···················· |
| Q | 4293 | hCoV-19/USA/CA-ALSR-2669/2020\|EPI_ISL_635620\|2020- | ···················· | ·A·················· | ···················· | ···················· | ···················· | ···················· |
| Q | 4294 | hCoV-19/USA/CA-ALSR-2670/2020\|EPI_ISL_635621\|2020- | ···················· | ·A·················· | ···················· | ···················· | ···················· | ···················· |
| Q | 4295 | hCoV-19/USA/CA-ALSR-2673/2020\|EPI_ISL_635622\|2020- | ···················· | ···················· | ···················· | ···················· | ···················· | ···················· |
| Q | 4296 | hCoV-19/USA/CA-ALSR-2674/2020\|EPI_ISL_635623\|2020- | ···················· | ···················· | ···················· | ···················· | ···················· | ···················· |
| Q | 4297 | hCoV-19/USA/CA-ALSR-2675/2020\|EPI_ISL_635624\|2020- | ···················· | ·A·················· | ···················· | ···················· | ···················· | ···················· |
| Q | 4298 | hCoV-19/USA/CA-ALSR-2676/2020\|EPI_ISL_635625\|2020- | ···················· | ·A·················· | ···················· | ···················· | ···················· | ···················· |
| Q | 4299 | hCoV-19/USA/CA-ALSR-2679/2020\|EPI_ISL_635626\|2020- | ···················· | ·A·················· | ···················· | ···················· | ···················· | ···················· |
| Q | 4300 | hCoV-19/USA/CA-ALSR-2681/2020\|EPI_ISL_635627\|2020- | ···················· | ···················· | ···················· | ···················· | ···················· | ···················· |
| Q | 4301 | hCoV-19/USA/CA-ALSR-2682/2020\|EPI_ISL_635628\|2020- | ···················· | ·A·················· | ···················· | ···················· | ···················· | ···················· |
| Q | 4302 | hCoV-19/USA/CA-ALSR-2684/2020\|EPI_ISL_635629\|2020- | ···················· | ·A·················· | ···················· | ···················· | ···················· | ···················· |
| Q | 4303 | hCoV-19/USA/CA-ALSR-2685/2020\|EPI_ISL_635630\|2020- | ···················· | ···················· | ···················· | ···················· | ···················· | ···················· |
| Q | 4304 | hCoV-19/USA/CA-ALSR-2689/2020\|EPI_ISL_635632\|2020- | ···················· | ·A·················· | ···················· | ···················· | ···················· | ···················· |
| Q | 4305 | hCoV-19/USA/CA-ALSR-2690/2020\|EPI_ISL_635633\|2020- | ···················· | ·A·················· | ···················· | ···················· | ···················· | ···················· |
| Q | 4306 | hCoV-19/USA/CA-ALSR-2691/2020\|EPI_ISL_635634\|2020- | ···················· | ···················· | ···················· | ···················· | ···················· | ···················· |
| Q | 4307 | hCoV-19/USA/CA-ALSR-2692/2020\|EPI_ISL_635635\|2020- | ···················· | ···················· | ···················· | ···················· | ···················· | ···················· |
| Q | 4308 | hCoV-19/USA/CA-ALSR-2697/2020\|EPI_ISL_635636\|2020- | ···················· | ·A·················· | ···················· | ···················· | ···················· | ···················· |
| Q | 4309 | hCoV-19/USA/CA-ALSR-2698/2020\|EPI_ISL_635637\|2020- | ···················· | ···················· | ···················· | ···················· | ···················· | ···················· |
| Q | 4310 | hCoV-19/USA/CA-ALSR-2703/2020\|EPI_ISL_635638\|2020- | ···················· | ·A·················· | ···················· | ···················· | ···················· | ···················· |
| Q | 4311 | hCoV-19/USA/CA-ALSR-2707/2020\|EPI_ISL_635639\|2020- | ···················· | ·A·················· | ···················· | ···················· | ···················· | ···················· |
| Q | 4312 | hCoV-19/USA/CA-ALSR-2711/2020\|EPI_ISL_635640\|2020- | ···················· | ···················· | ···················· | ···················· | ···················· | ···················· |
| Q | 4313 | hCoV-19/USA/CA-ALSR-2714/2020\|EPI_ISL_635642\|2020- | ···················· | ···················· | ···················· | ···················· | ···················· | ···················· |
| Q | 4314 | hCoV-19/USA/CA-ALSR-2716/2020\|EPI_ISL_635643\|2020- | ···················· | ·A·················· | ···················· | ···················· | ···················· | ···················· |
| Q | 4315 | hCoV-19/USA/CA-ALSR-2722/2020\|EPI_ISL_635644\|2020- | ···················· | ·A·················· | ···················· | ···················· | ···················· | ···················· |
| Q | 4316 | hCoV-19/USA/CA-ALSR-2727/2020\|EPI_ISL_635646\|2020- | ···················· | ·A·················· | ···················· | ···················· | ···················· | ···················· |
| Q | 4317 | hCoV-19/USA/CA-ALSR-2775/2020\|EPI_ISL_635673\|2020- | ···················· | ···················· | ···················· | ···················· | ···················· | ···················· |
| Q | 4318 | hCoV-19/USA/CA-ALSR-2776/2020\|EPI_ISL_635674\|2020- | ···················· | ·A·················· | ···················· | ···················· | ···················· | ···················· |
| Q | 4320 | hCoV-19/Canada/ON-PHL-20-01786/2020\|EPI_ISL_636936 | ···················· | ···················· | ···················· | ···················· | ···················· | ···················· |
| Q | 4321 | hCoV-19/Canada/ON-PHL-20-01787/2020\|EPI_ISL_636937 | ···················· | ······A············· | ···················· | ···················· | ···················· | ···················· |
| Q | 4322 | hCoV-19/Canada/ON-PHL-20-01788/2020\|EPI_ISL_636938 | ···················· | ······A············· | ···················· | ···················· | ···················· | ···················· |
| Q | 4324 | hCoV-19/Canada/ON-PHL-20-01790/2020\|EPI_ISL_636940 | ···················· | ···················· | ···················· | ···················· | ···················· | ···················· |
| Q | 4325 | hCoV-19/Canada/ON-PHL-20-01791/2020\|EPI_ISL_636941 | ···················· | ···················· | ···················· | ···················· | ···················· | ···················· |
| Q | 4328 | hCoV-19/Canada/ON-PHL-20-01794/2020\|EPI_ISL_636944 | ···················· | ···················· | ···················· | ···················· | ···················· | ···················· |
| Q | 4330 | hCoV-19/Canada/ON-PHL-20-01796/2020\|EPI_ISL_636946 | ···················· | ···················· | ···················· | ···················· | ···················· | ···················· |
| Q | 4331 | hCoV-19/Canada/ON-PHL-20-01797/2020\|EPI_ISL_636947 | ···················· | ···················· | ···················· | ···················· | ···················· | ···················· |
| Q | 4333 | hCoV-19/Canada/ON-PHL-20-01806/2020\|EPI_ISL_636950 | ···················· | ···················· | ···················· | ···················· | ···················· | ···················· |
| Q | 4334 | hCoV-19/Canada/ON-PHL-20-01807/2020\|EPI_ISL_636951 | ···················· | ···················· | ···················· | ···················· | ···················· | ···················· |
| Q | 4335 | hCoV-19/Canada/ON-PHL-20-01808/2020\|EPI_ISL_636952 | ···················· | ···················· | ···················· | ···················· | ···················· | ···················· |
| Q | 4336 | hCoV-19/Canada/ON-PHL-20-01809/2020\|EPI_ISL_636953 | ···················· | ···················· | ···················· | ···················· | ···················· | ···················· |
| Q | 4338 | hCoV-19/Canada/ON-PHL-20-01811/2020\|EPI_ISL_636955 | ···················· | ···················· | ···················· | ···················· | ···················· | ···················· |
| Q | 4340 | hCoV-19/Canada/ON-PHL-20-01813/2020\|EPI_ISL_636957 | ···················· | ···················· | ···················· | ···················· | ···················· | ···················· |
| Q | 4341 | hCoV-19/Canada/ON-PHL-20-01814/2020\|EPI_ISL_636958 | ···················· | ···················· | ···················· | ···················· | ···················· | ···················· |
| Q | 4342 | hCoV-19/Canada/ON-PHL-20-01815/2020\|EPI_ISL_636959 | ···················· | ···················· | ···················· | ···················· | ···················· | ···················· |
| Q | 4343 | hCoV-19/Saudi Arabia/KAUST-MADINAH893/2020\|EPI_ISL | ···················· | ···················· | ···················· | ···················· | ···················· | ···················· |
| Q | 4344 | hCoV-19/Saudi Arabia/KAUST-JEDDAH994/2020\|EPI_ISL_ | ···················· | ···················· | ···················· | ···················· | ···················· | ···················· |
| Q | 4345 | hCoV-19/Saudi Arabia/KAUST-JEDDAH1056/2020\|EPI_ISL | ···················· | ···················· | ···················· | ···················· | ···················· | ···················· |
| Q | 4346 | hCoV-19/Saudi Arabia/KAUST-MADINAH1080/2020\|EPI_IS | ···················· | ···················· | ···················· | ···················· | ···················· | ···················· |
| Q | 4347 | hCoV-19/Turkey/ETLKVET1/2020\|EPI_ISL_636969\|2020-0 | ···················· | ···················· | ···················· | ···················· | ···················· | ···················· |
| Q | 4348 | hCoV-19/Turkey/ETLKVET2/2020\|EPI_ISL_636970\|2020-0 | ···················· | ···················· | ···················· | ···················· | ···················· | ···················· |
| Q | 4349 | hCoV-19/Turkey/ETLKVET3/2020\|EPI_ISL_636971\|2020-0 | ···················· | ···················· | ···················· | ···················· | ···················· | ···················· |
| Q | 4350 | hCoV-19/Saudi Arabia/KAUST-jeddah839/2020\|EPI_ISL_ | ···················· | ···················· | ···················· | ···················· | ···················· | ···················· |
| Q | 4351 | hCoV-19/Bahrain/023341084/2020\|EPI_ISL_636973\|2020 | ···················· | ···················· | ···················· | ···················· | ···················· | ···················· |
| Q | 4352 | hCoV-19/Mozambique/KRISP-MZ157469/2020\|EPI_ISL_636 | ···················· | ···················· | ···················· | ···················· | ···················· | ···················· |
| Q | 4353 | hCoV-19/Northern Ireland/NIRE-108B60/2020\|EPI_ISL_ | ···················· | ···················· | ···················· | ···················· | ···················· | ···················· |
| Q | 4354 | hCoV-19/England/NORW-F21B3/2020\|EPI_ISL_627071\|202 | ···················· | ···················· | ···················· | ···················· | ···················· | ···················· |
| Q | 4356 | hCoV-19/England/NORW-F2344/2020\|EPI_ISL_627073\|202 | ···················· | ···················· | ···················· | ···················· | ···················· | ···················· |

| | | | | | | | |
|---|---|---|---|---|---|---|---|
| Q | 4359 hCoV-19/England/NORW-F36C2/2020\|EPI_ISL_627076\|202 | ···················· | ···················· | ····················· | ···················· | ···················· | ···················· |
| Q | 4360 hCoV-19/England/NORW-F3DCD/2020\|EPI_ISL_627077\|202 | ···················· | ···················· | ····················· | ···················· | ···················· | ···················· |
| Q | 4361 hCoV-19/Wales/PHWC-483CC1/2020\|EPI_ISL_627078\|2020 | ···················· | ···················· | ····················· | ···················· | ···················· | ···················· |
| Q | 4362 hCoV-19/England/QEUH-A4D1FB/2020\|EPI_ISL_627079\|20 | ···················· | ·A·················· | ····················· | ···················· | ···················· | ···················· |
| Q | 4363 hCoV-19/England/NORW-F16E2/2020\|EPI_ISL_627080\|202 | ···················· | ···················· | ····················· | ···················· | ···················· | ···················· |
| Q | 4364 hCoV-19/England/NORW-F1BF2/2020\|EPI_ISL_627081\|202 | ·······T············ | ···················· | ····················· | ···················· | ···················· | ···················· |
| Q | 4365 hCoV-19/England/LOND-1262E85/2020\|EPI_ISL_627082\|2 | ···················· | ·A·················· | ····················· | ···················· | ···················· | ··A················· |
| Q | 4366 hCoV-19/England/NORW-F21D1/2020\|EPI_ISL_627083\|202 | ···················· | ·A·················· | ····················· | ···················· | ···················· | ··A················· |
| Q | 4367 hCoV-19/England/NORW-F2423/2020\|EPI_ISL_627084\|202 | ···················· | ·A·················· | ····················· | ···················· | ···················· | ··A················· |
| Q | 4368 hCoV-19/England/QEUH-A49872/2020\|EPI_ISL_627085\|20 | ···················· | ·A·················· | ····················· | ···················· | ···················· | ··A················· |
| Q | 4369 hCoV-19/Wales/PHWC-483F13/2020\|EPI_ISL_627086\|2020 | ···················· | ···················· | ····················· | ···················· | ···················· | ···················· |
| Q | 4370 hCoV-19/Wales/PHWC-4858E9/2020\|EPI_ISL_627087\|2020 | ···················· | ···················· | ····················· | ···················· | ···················· | ···················· |
| Q | 4371 hCoV-19/Wales/PHWC-483E34/2020\|EPI_ISL_627088\|2020 | ···················· | ···················· | ····················· | ···················· | ···················· | ···················· |
| Q | 4372 hCoV-19/Wales/PHWC-483F40/2020\|EPI_ISL_627089\|2020 | ···················· | ···················· | ····················· | ···················· | ···················· | ···················· |
| Q | 4373 hCoV-19/Wales/PHWC-484264/2020\|EPI_ISL_627090\|2020 | ···················· | ···················· | ····················· | ···················· | ···················· | ···················· |
| Q | 4374 hCoV-19/Wales/PHWC-48454D/2020\|EPI_ISL_627091\|2020 | ···················· | ···················· | ····················· | ···················· | ···················· | ···················· |
| Q | 4375 hCoV-19/Wales/PHWC-48456B/2020\|EPI_ISL_627092\|2020 | ···················· | ···················· | ····················· | ···················· | ···················· | ···················· |
| Q | 4376 hCoV-19/Wales/PHWC-4849D8/2020\|EPI_ISL_627093\|2020 | ···················· | ···················· | ····················· | ···················· | ···················· | ···················· |
| Q | 4377 hCoV-19/Wales/PHWC-484C1B/2020\|EPI_ISL_627094\|2020 | ···················· | ···················· | ····················· | ···················· | ···················· | ···················· |
| Q | 4378 hCoV-19/Wales/PHWC-484CDF/2020\|EPI_ISL_627095\|2020 | ···················· | ···················· | ····················· | ···················· | ···················· | ···················· |
| Q | 4379 hCoV-19/Wales/PHWC-485F11/2020\|EPI_ISL_627096\|2020 | ···················· | ···················· | ····················· | ···················· | ···················· | ···················· |
| Q | 4380 hCoV-19/Wales/PHWC-4831D1/2020\|EPI_ISL_627097\|2020 | ···················· | ···················· | ····················· | ···················· | ···················· | ···················· |
| Q | 4381 hCoV-19/England/QEUH-A3285E/2020\|EPI_ISL_628360\|20 | ···················· | ···················· | ····················· | ···················· | ···················· | ···················· |
| Q | 4382 hCoV-19/England/QEUH-A3286D/2020\|EPI_ISL_628361\|20 | ···················· | ···················· | ····················· | ···················· | ···················· | ···················· |
| Q | 4383 hCoV-19/England/QEUH-A3287C/2020\|EPI_ISL_628362\|20 | ···················· | ···················· | ····················· | ···················· | ···················· | ···················· |
| Q | 4384 hCoV-19/England/QEUH-A3289A/2020\|EPI_ISL_628363\|20 | ···················· | ·A·················· | ····················· | ···················· | ···················· | ··A················· |
| Q | 4385 hCoV-19/England/QEUH-A328D6/2020\|EPI_ISL_628364\|20 | ···················· | ···················· | ····················· | ···················· | ···················· | ···················· |
| Q | 4386 hCoV-19/England/QEUH-A328E5/2020\|EPI_ISL_628365\|20 | ···················· | ···················· | ····················· | ···················· | ···················· | ···················· |
| Q | 4387 hCoV-19/England/QEUH-A328F4/2020\|EPI_ISL_628366\|20 | ···················· | ···················· | ····················· | ···················· | ···················· | ···················· |
| Q | 4388 hCoV-19/England/QEUH-A3291F/2020\|EPI_ISL_628367\|20 | ···················· | ···················· | ····················· | ···················· | ···················· | ···················· |
| Q | 4389 hCoV-19/England/QEUH-A3293D/2020\|EPI_ISL_628368\|20 | ···················· | ···················· | ····················· | ···················· | ···················· | ···················· |
| Q | 4390 hCoV-19/England/QEUH-A3326E/2020\|EPI_ISL_628369\|20 | ···················· | ···················· | ····················· | ···················· | ···················· | ···················· |
| Q | 4391 hCoV-19/England/QEUH-A332B9/2020\|EPI_ISL_628370\|20 | ···················· | ···················· | ····················· | ···················· | ···················· | ···················· |
| Q | 4392 hCoV-19/England/QEUH-A332C8/2020\|EPI_ISL_628371\|20 | ···················· | ···················· | ····················· | ···················· | ···················· | ···················· |
| Q | 4393 hCoV-19/England/QEUH-A3335C/2020\|EPI_ISL_628372\|20 | ···················· | ···················· | ····················· | ···················· | ···················· | ···················· |
| Q | 4394 hCoV-19/England/QEUH-A3336B/2020\|EPI_ISL_628373\|20 | ···················· | ···················· | ····················· | ···················· | ···················· | ···················· |
| Q | 4395 hCoV-19/England/QEUH-A333A7/2020\|EPI_ISL_628374\|20 | ···················· | ···················· | ····················· | ···················· | ···················· | ···················· |
| Q | 4396 hCoV-19/England/QEUH-A333E3/2020\|EPI_ISL_628375\|20 | ···················· | ···················· | ····················· | ···················· | ···················· | ···················· |
| Q | 4397 hCoV-19/England/QEUH-A333F2/2020\|EPI_ISL_628376\|20 | ···················· | ···················· | ····················· | ···················· | ···················· | ···················· |
| Q | 4398 hCoV-19/England/QEUH-A3340E/2020\|EPI_ISL_628377\|20 | ···················· | ···················· | ····················· | ···················· | ···················· | ···················· |
| Q | 4399 hCoV-19/England/QEUH-A3341D/2020\|EPI_ISL_628378\|20 | ···················· | ···················· | ····················· | ···················· | ···················· | ···················· |
| Q | 4400 hCoV-19/England/QEUH-A3342C/2020\|EPI_ISL_628379\|20 | ···················· | ···················· | ····················· | ···················· | ···················· | ···················· |
| Q | 4401 hCoV-19/England/QEUH-A3343B/2020\|EPI_ISL_628380\|20 | ···················· | ···················· | ····················· | ···················· | ···················· | ···················· |
| Q | 4402 hCoV-19/England/QEUH-A33468/2020\|EPI_ISL_628381\|20 | ···················· | ···················· | ····················· | ···················· | ···················· | ···················· |
| Q | 4403 hCoV-19/England/QEUH-A33477/2020\|EPI_ISL_628382\|20 | ···················· | ···················· | ····················· | ···················· | ···················· | ···················· |
| Q | 4404 hCoV-19/England/QEUH-A334A4/2020\|EPI_ISL_628383\|20 | ···················· | ···················· | ····················· | ···················· | ···················· | ···················· |
| Q | 4405 hCoV-19/England/QEUH-A334B3/2020\|EPI_ISL_628384\|20 | ···················· | ···················· | ····················· | ···················· | ···················· | ···················· |
| Q | 4406 hCoV-19/England/QEUH-A334C2/2020\|EPI_ISL_628385\|20 | ···················· | ···················· | ····················· | ···················· | ···················· | ···················· |
| Q | 4407 hCoV-19/England/QEUH-A334E0/2020\|EPI_ISL_628386\|20 | ···················· | ···················· | ····················· | ···················· | ···················· | ···················· |
| Q | 4408 hCoV-19/England/QEUH-A334FF/2020\|EPI_ISL_628387\|20 | ···················· | ···················· | ····················· | ···················· | ···················· | ···················· |
| Q | 4409 hCoV-19/England/QEUH-A3350B/2020\|EPI_ISL_628388\|20 | ···················· | ···················· | ····················· | ···················· | ···················· | ···················· |
| Q | 4410 hCoV-19/Scotland/QEUH-A3351A/2020\|EPI_ISL_628389\|2 | ···················· | ···················· | ····················· | ···················· | ···················· | ···················· |
| Q | 4411 hCoV-19/England/QEUH-A33529/2020\|EPI_ISL_628390\|20 | ···················· | ···················· | ····················· | ···················· | ···················· | ···················· |
| Q | 4412 hCoV-19/England/QEUH-A33538/2020\|EPI_ISL_628391\|20 | ···················· | ···················· | ····················· | ···················· | ···················· | ···················· |
| Q | 4413 hCoV-19/England/QEUH-A33547/2020\|EPI_ISL_628392\|20 | ···················· | ···················· | ····················· | ···················· | ···················· | ···················· |
| Q | 4414 hCoV-19/England/QEUH-A33556/2020\|EPI_ISL_628393\|20 | ···················· | ···················· | ····················· | ···················· | ···················· | ···················· |
| Q | 4415 hCoV-19/Wales/QEUH-A33592/2020\|EPI_ISL_628394\|2020 | ···················· | ···················· | ····················· | ···················· | ···················· | ···················· |
| Q | 4416 hCoV-19/England/QEUH-A335B0/2020\|EPI_ISL_628395\|20 | ···················· | ···················· | ····················· | ···················· | ···················· | ···················· |
| Q | 4417 hCoV-19/England/QEUH-A335ED/2020\|EPI_ISL_628396\|20 | ···················· | ···················· | ····················· | ···················· | ···················· | ···················· |
| Q | 4418 hCoV-19/Scotland/QEUH-A33635/2020\|EPI_ISL_628397\|2 | ···················· | ···················· | ····················· | ···················· | ···················· | ···················· |
| Q | 4419 hCoV-19/Scotland/QEUH-A33644/2020\|EPI_ISL_628398\|2 | ···················· | ···················· | ····················· | ···················· | ···················· | ···················· |
| Q | 4420 hCoV-19/Scotland/QEUH-A33671/2020\|EPI_ISL_628399\|2 | ···················· | ···················· | ····················· | ···················· | ···················· | ···················· |
| Q | 4421 hCoV-19/Scotland/QEUH-A33680/2020\|EPI_ISL_628400\|2 | ···················· | ···················· | ····················· | ···················· | ···················· | ···················· |
| Q | 4422 hCoV-19/England/QEUH-A336AE/2020\|EPI_ISL_628401\|20 | ···················· | ···················· | ····················· | ···················· | ···················· | ···················· |
| Q | 4423 hCoV-19/England/QEUH-A336BD/2020\|EPI_ISL_628402\|20 | ···················· | ···················· | ····················· | ···················· | ···················· | ···················· |
| Q | 4424 hCoV-19/England/QEUH-A336CC/2020\|EPI_ISL_628403\|20 | ···················· | ···················· | ····················· | ···················· | ···················· | ···················· |
| Q | 4425 hCoV-19/England/QEUH-A336EA/2020\|EPI_ISL_628404\|20 | ···················· | ···················· | ····················· | ···················· | ···················· | ···················· |
| Q | 4426 hCoV-19/England/QEUH-A336F9/2020\|EPI_ISL_628405\|20 | ···················· | ···················· | ····················· | ···················· | ···················· | ···················· |
| Q | 4427 hCoV-19/England/QEUH-A33705/2020\|EPI_ISL_628406\|20 | ···················· | ···················· | ····················· | ···················· | ···················· | ···················· |
| Q | 4428 hCoV-19/England/QEUH-A33732/2020\|EPI_ISL_628407\|20 | ···················· | ···················· | ····················· | ···················· | ···················· | ···················· |
| Q | 4429 hCoV-19/England/QEUH-A337AB/2020\|EPI_ISL_628408\|20 | ···················· | ···················· | ····················· | ···················· | ···················· | ···················· |
| Q | 4430 hCoV-19/England/QEUH-A337E7/2020\|EPI_ISL_628409\|20 | ···················· | ···················· | ····················· | ···················· | ······T············· | ···················· |
| Q | 4431 hCoV-19/England/QEUH-A33811/2020\|EPI_ISL_628410\|20 | ···················· | ···················· | ····················· | ···················· | ···················· | ···················· |
| Q | 4432 hCoV-19/England/QEUH-A3386C/2020\|EPI_ISL_628411\|20 | ···················· | ···················· | ····················· | ···················· | ···················· | ···················· |
| Q | 4433 hCoV-19/England/QEUH-A33899/2020\|EPI_ISL_628412\|20 | ···················· | ···················· | ····················· | ···················· | ···················· | ···················· |
| Q | 4434 hCoV-19/England/QEUH-A338B7/2020\|EPI_ISL_628413\|20 | ···················· | ···················· | ····················· | ···················· | ···················· | ···················· |
| Q | 4435 hCoV-19/England/QEUH-A338C6/2020\|EPI_ISL_628414\|20 | ···················· | ···················· | ····················· | ···················· | ···················· | ···················· |
| Q | 4436 hCoV-19/Wales/QEUH-A3392D/2020\|EPI_ISL_628415\|2020 | ···················· | ···················· | ····················· | ···················· | ···················· | ···················· |
| Q | 4437 hCoV-19/England/QEUH-A3395A/2020\|EPI_ISL_628416\|20 | ···················· | ···················· | ····················· | ···················· | ···················· | ···················· |
| Q | 4438 hCoV-19/England/QEUH-A33969/2020\|EPI_ISL_628417\|20 | ···················· | ···················· | ····················· | ···················· | ···················· | ···················· |

| | | | | | | | | |
|---|---|---|---|---|---|---|---|---|
| Q | 4439 | hCoV-19/England/QEUH-A33978/2020\|EPI_ISL_628418\|20 | ···················· | ···················· | ····················· | ···················· | ···················· | ···················· |
| Q | 4440 | hCoV-19/England/QEUH-A33996/2020\|EPI_ISL_628419\|20 | ···················· | ···················· | ····················· | ···················· | ···················· | ···················· |
| Q | 4441 | hCoV-19/Wales/QEUH-A339A5/2020\|EPI_ISL_628420\|2020 | ···················· | ···················· | ····················· | ···················· | ···················· | ···················· |
| Q | 4442 | hCoV-19/England/QEUH-A341D9/2020\|EPI_ISL_628421\|20 | ···················· | ···················· | ····················· | ···················· | ···················· | ···················· |
| Q | 4443 | hCoV-19/England/QEUH-A34EB4/2020\|EPI_ISL_628422\|20 | ···················· | ···················· | ····················· | ···················· | ···················· | ···················· |
| Q | 4444 | hCoV-19/England/QEUH-A34EC3/2020\|EPI_ISL_628423\|20 | ···················· | ·A·················· | ····················· | ···················· | ···················· | ··A················· |
| Q | 4445 | hCoV-19/England/QEUH-A34EE1/2020\|EPI_ISL_628424\|20 | ···················· | ···················· | ····················· | ···················· | ···················· | ···················· |
| Q | 4446 | hCoV-19/England/QEUH-A34EF0/2020\|EPI_ISL_628425\|20 | ···················· | ···················· | ····················· | ···················· | ···················· | ···················· |
| Q | 4447 | hCoV-19/England/QEUH-A34F1B/2020\|EPI_ISL_628426\|20 | ···················· | ···················· | ····················· | ···················· | ···················· | ···················· |
| Q | 4448 | hCoV-19/England/QEUH-A34F2A/2020\|EPI_ISL_628427\|20 | ···················· | ···················· | ····················· | ···················· | ···················· | ···················· |
| Q | 4449 | hCoV-19/England/QEUH-A34F39/2020\|EPI_ISL_628428\|20 | ···················· | ···················· | ····················· | ···················· | ···················· | ···················· |
| Q | 4450 | hCoV-19/Scotland/QEUH-A4963C/2020\|EPI_ISL_628429\|2 | ···················· | ···················· | ····················· | ···················· | ···················· | ···················· |
| Q | 4451 | hCoV-19/England/QEUH-A497C0/2020\|EPI_ISL_628430\|20 | ···················· | ···················· | ····················· | ···················· | ···················· | ···················· |
| Q | 4452 | hCoV-19/England/QEUH-A49809/2020\|EPI_ISL_628431\|20 | ···················· | ···················· | ····················· | ···················· | ···················· | ···················· |
| Q | 4453 | hCoV-19/England/QEUH-A49818/2020\|EPI_ISL_628432\|20 | ···················· | ···················· | ····················· | ···················· | ···················· | ···················· |
| Q | 4454 | hCoV-19/England/QEUH-A49827/2020\|EPI_ISL_628433\|20 | ···················· | ···················· | ····················· | ···················· | ···················· | ···················· |
| Q | 4455 | hCoV-19/England/QEUH-A49836/2020\|EPI_ISL_628434\|20 | ···················· | ···················· | ····················· | ···················· | ·····T·············· | ···················· |
| Q | 4456 | hCoV-19/England/QEUH-A49863/2020\|EPI_ISL_628435\|20 | ···················· | ···················· | ····················· | ···················· | ···················· | ···················· |
| Q | 4457 | hCoV-19/Scotland/QEUH-A498BE/2020\|EPI_ISL_628436\|2 | ···················· | ···················· | ····················· | ···················· | ···················· | ···················· |
| Q | 4458 | hCoV-19/England/QEUH-A49906/2020\|EPI_ISL_628437\|20 | ···················· | ···················· | ····················· | ···················· | ···················· | ···················· |
| Q | 4459 | hCoV-19/England/QEUH-A49924/2020\|EPI_ISL_628438\|20 | ···················· | ···················· | ····················· | ···················· | ···················· | ···················· |
| Q | 4460 | hCoV-19/England/QEUH-A49951/2020\|EPI_ISL_628439\|20 | ···················· | ···················· | ····················· | ···················· | ···················· | ···················· |
| Q | 4461 | hCoV-19/England/QEUH-A4AF03/2020\|EPI_ISL_628460\|20 | ···················· | ···················· | ····················· | ···················· | ···················· | ···················· |
| Q | 4462 | hCoV-19/Scotland/QEUH-A4AF4F/2020\|EPI_ISL_628461\|2 | ···················· | ···················· | ····················· | ···················· | ···················· | ···················· |
| Q | 4463 | hCoV-19/Scotland/QEUH-A4AF5E/2020\|EPI_ISL_628462\|2 | ···················· | ···················· | ····················· | ···················· | ···················· | ···················· |
| Q | 4464 | hCoV-19/Wales/QEUH-A4AFB8/2020\|EPI_ISL_628463\|2020 | ···················· | ···················· | ····················· | ···················· | ···················· | ···················· |
| Q | 4465 | hCoV-19/England/QEUH-A4AFC7/2020\|EPI_ISL_628464\|20 | ···················· | ···················· | ····················· | ···················· | ···················· | ···················· |
| Q | 4466 | hCoV-19/Scotland/QEUH-A4B315/2020\|EPI_ISL_628465\|2 | ···················· | ···················· | ····················· | ···················· | ···················· | ···················· |
| Q | 4467 | hCoV-19/Scotland/QEUH-A4B324/2020\|EPI_ISL_628466\|2 | ···················· | ···················· | ····················· | ···················· | ···················· | ···················· |
| Q | 4468 | hCoV-19/Scotland/QEUH-A4B38E/2020\|EPI_ISL_628467\|2 | ···················· | ···················· | ····················· | ···················· | ···················· | ···················· |
| Q | 4469 | hCoV-19/Scotland/QEUH-A4B3AC/2020\|EPI_ISL_628468\|2 | ···················· | ···················· | ····················· | ···················· | ···················· | ···················· |
| Q | 4470 | hCoV-19/Scotland/QEUH-A4B3BB/2020\|EPI_ISL_628469\|2 | ···················· | ···················· | ····················· | ···················· | ···················· | ···················· |
| Q | 4471 | hCoV-19/Scotland/QEUH-A4B658/2020\|EPI_ISL_628470\|2 | ···················· | ···················· | ····················· | ···················· | ···················· | ···················· |
| Q | 4472 | hCoV-19/Scotland/QEUH-A4B6B2/2020\|EPI_ISL_628471\|2 | ···················· | ···················· | ····················· | ···················· | ···················· | ···················· |
| Q | 4473 | hCoV-19/Scotland/QEUH-A4B6D0/2020\|EPI_ISL_628472\|2 | ···················· | ···················· | ····················· | ···················· | ···················· | ···················· |
| Q | 4474 | hCoV-19/Scotland/QEUH-A4B6FE/2020\|EPI_ISL_628473\|2 | ···················· | ···················· | ····················· | ···················· | ···················· | ···················· |
| Q | 4475 | hCoV-19/Scotland/QEUH-A4B728/2020\|EPI_ISL_628474\|2 | ···················· | ···················· | ····················· | ···················· | ···················· | ···················· |
| Q | 4476 | hCoV-19/Scotland/QEUH-A4B737/2020\|EPI_ISL_628475\|2 | ···················· | ···················· | ····················· | ···················· | ···················· | ···················· |
| Q | 4477 | hCoV-19/Scotland/QEUH-A4B746/2020\|EPI_ISL_628476\|2 | ···················· | ···················· | ····················· | ···················· | ···················· | ···················· |
| Q | 4478 | hCoV-19/Scotland/QEUH-A4B773/2020\|EPI_ISL_628477\|2 | ···················· | ···················· | ····················· | ···················· | ···················· | ···················· |
| Q | 4479 | hCoV-19/Scotland/QEUH-A4B782/2020\|EPI_ISL_628478\|2 | ···················· | ···················· | ····················· | ···················· | ···················· | ···················· |
| Q | 4480 | hCoV-19/Scotland/QEUH-A4B7BF/2020\|EPI_ISL_628479\|2 | ···················· | ···················· | ····················· | ···················· | ···················· | ···················· |
| Q | 4481 | hCoV-19/England/MILK-AC90AF/2020\|EPI_ISL_629691\|20 | ···················· | ···················· | ····················· | ···················· | ···················· | ···················· |
| Q | 4482 | hCoV-19/England/MILK-AC8538/2020\|EPI_ISL_629693\|20 | ···················· | ···················· | ····················· | ···················· | ···················· | ···················· |
| Q | 4483 | hCoV-19/England/MILK-AC90CD/2020\|EPI_ISL_629694\|20 | ···················· | ···················· | ····················· | ···················· | ···················· | ···················· |
| Q | 4484 | hCoV-19/England/MILK-AC8FEF/2020\|EPI_ISL_629695\|20 | ···················· | ···················· | ····················· | ···················· | ···················· | ···················· |
| Q | 4485 | hCoV-19/England/MILK-ACE5AB/2020\|EPI_ISL_629696\|20 | ···················· | ···················· | ····················· | ···················· | ···················· | ···················· |
| Q | 4486 | hCoV-19/England/MILK-AC7F4A/2020\|EPI_ISL_629697\|20 | ···················· | ···················· | ····················· | ···················· | ···················· | ···················· |
| Q | 4487 | hCoV-19/England/MILK-AC8E5B/2020\|EPI_ISL_629698\|20 | ···················· | ···················· | ····················· | ···················· | ···················· | ···················· |
| Q | 4488 | hCoV-19/England/MILK-ACE523/2020\|EPI_ISL_629730\|20 | ···················· | ···················· | ····················· | ···················· | ···················· | ···················· |
| Q | 4489 | hCoV-19/England/MILK-ACE9AF/2020\|EPI_ISL_629731\|20 | ···················· | ···················· | ····················· | ···················· | ···················· | ···················· |
| Q | 4490 | hCoV-19/England/MILK-AC8064/2020\|EPI_ISL_629732\|20 | ···················· | ···················· | ····················· | ···················· | ···················· | ···················· |
| Q | 4491 | hCoV-19/England/MILK-AC8B63/2020\|EPI_ISL_629734\|20 | ···················· | ···················· | ····················· | ···················· | ·····T·············· | ···················· |
| Q | 4492 | hCoV-19/England/MILK-ACEE0A/2020\|EPI_ISL_629737\|20 | ···················· | ···················· | ····················· | ···················· | ···················· | ···················· |
| Q | 4493 | hCoV-19/England/MILK-ACEF34/2020\|EPI_ISL_629738\|20 | ···················· | ···················· | ····················· | ···················· | ·····T·············· | ···················· |
| Q | 4494 | hCoV-19/England/MILK-ACF267/2020\|EPI_ISL_629740\|20 | ···················· | ···················· | ····················· | ···················· | ···················· | ···················· |
| Q | 4495 | hCoV-19/England/MILK-ACF7B3/2020\|EPI_ISL_629743\|20 | ···················· | ···················· | ····················· | ···················· | ·····T·············· | ···················· |
| Q | 4496 | hCoV-19/England/MILK-AC9230/2020\|EPI_ISL_629744\|20 | ···················· | ···················· | ····················· | ···················· | ···················· | ···················· |
| Q | 4497 | hCoV-19/England/MILK-ACEB03/2020\|EPI_ISL_629745\|20 | ···················· | ···················· | ····················· | ···················· | ·····T·············· | ···················· |
| Q | 4498 | hCoV-19/England/MILK-ACEA33/2020\|EPI_ISL_629746\|20 | ···················· | ·A·················· | ····················· | ···················· | ···················· | ···················· |
| Q | 4499 | hCoV-19/England/MILK-AC892D/2020\|EPI_ISL_629748\|20 | ···················· | ···················· | ····················· | ···················· | ···················· | ···················· |
| Q | 4500 | hCoV-19/England/MILK-ACEA42/2020\|EPI_ISL_629749\|20 | ···················· | ···················· | ····················· | ···················· | ···················· | ···················· |
| Q | 4501 | hCoV-19/England/MILK-ACEDEF/2020\|EPI_ISL_629750\|20 | ···················· | ···················· | ····················· | ···················· | ···················· | ···················· |
| Q | 4502 | hCoV-19/England/MILK-AC8F2B/2020\|EPI_ISL_629751\|20 | ···················· | ···················· | ····················· | ···················· | ···················· | ···················· |
| Q | 4503 | hCoV-19/England/MILK-ACF2EF/2020\|EPI_ISL_629752\|20 | ···················· | ···················· | ····················· | ···················· | ···················· | ···················· |
| Q | 4504 | hCoV-19/England/QEUH-AD0360/2020\|EPI_ISL_629755\|20 | ···················· | ···················· | ····················· | ···················· | ···················· | ···················· |
| Q | 4505 | hCoV-19/England/QEUH-AD3DAB/2020\|EPI_ISL_629756\|20 | ···················· | ···················· | ····················· | ···················· | ···················· | ···················· |
| Q | 4506 | hCoV-19/England/QEUH-AD3D7E/2020\|EPI_ISL_629757\|20 | ···················· | ···················· | ····················· | ···················· | ···················· | ···················· |
| Q | 4507 | hCoV-19/England/QEUH-AD3AFF/2020\|EPI_ISL_629758\|20 | ···················· | ···················· | ····················· | ···················· | ···················· | ···················· |
| Q | 4508 | hCoV-19/England/QEUH-AD1D61/2020\|EPI_ISL_629759\|20 | ···················· | ···················· | ····················· | ···················· | ···················· | ···················· |
| Q | 4509 | hCoV-19/England/QEUH-AD6379/2020\|EPI_ISL_629760\|20 | ···················· | ···················· | ····················· | ···················· | ···················· | ···················· |
| Q | 4510 | hCoV-19/England/QEUH-AD260B/2020\|EPI_ISL_629761\|20 | ···················· | ···················· | ····················· | ···················· | ···················· | ···················· |
| Q | 4511 | hCoV-19/England/QEUH-AD09C8/2020\|EPI_ISL_629762\|20 | ···················· | ···················· | ····················· | ···················· | ···················· | ···················· |
| Q | 4512 | hCoV-19/England/QEUH-AD048B/2020\|EPI_ISL_629763\|20 | ···················· | ···················· | ····················· | ···················· | ·····T·············· | ···················· |
| Q | 4513 | hCoV-19/England/QEUH-AD063A/2020\|EPI_ISL_629764\|20 | ···················· | ···················· | ····················· | ···················· | ···················· | ···················· |
| Q | 4514 | hCoV-19/England/QEUH-AD03D9/2020\|EPI_ISL_629765\|20 | ···················· | ···················· | ····················· | ···················· | ···················· | ···················· |
| Q | 4515 | hCoV-19/England/QEUH-AD3E8A/2020\|EPI_ISL_629766\|20 | ···················· | ···················· | ····················· | ···················· | ···················· | ···················· |
| Q | 4516 | hCoV-19/England/QEUH-AD098C/2020\|EPI_ISL_629767\|20 | ···················· | ···················· | ····················· | ···················· | ···················· | ···················· |
| Q | 4517 | hCoV-19/England/QEUH-AD2559/2020\|EPI_ISL_629768\|20 | ···················· | ···················· | ····················· | ···················· | ···················· | ···················· |
| Q | 4518 | hCoV-19/England/QEUH-AD2780/2020\|EPI_ISL_629769\|20 | ···················· | ···················· | ····················· | ···················· | ···················· | ···················· |

| | | | | | | | |
|---|---|---|---|---|---|---|---|
| Q | 4519 hCoV-19/England/QEUH-AD25A4/2020\|EPI_ISL_629790\|20 | ···················· | ···················· | ···················· | ···················· | ···················· | ···················· |
| Q | 4520 hCoV-19/England/QEUH-AD06B2/2020\|EPI_ISL_629791\|20 | ···················· | ···················· | ···················· | ···················· | ···················· | ···················· |
| Q | 4521 hCoV-19/England/QEUH-AD0922/2020\|EPI_ISL_629792\|20 | ···················· | ···················· | ···················· | ···················· | ···················· | ···················· |
| Q | 4522 hCoV-19/England/QEUH-AD3D32/2020\|EPI_ISL_629793\|20 | ···················· | ···················· | ···················· | ···················· | ···················· | ···················· |
| Q | 4523 hCoV-19/England/QEUH-AD2F1F/2020\|EPI_ISL_629794\|20 | ···················· | ···················· | ···················· | ···················· | ···················· | ···················· |
| Q | 4524 hCoV-19/England/QEUH-AD02EB/2020\|EPI_ISL_629795\|20 | ···················· | ···················· | ···················· | ···················· | ·····T·············· | ···················· |
| Q | 4525 hCoV-19/England/QEUH-AD2E30/2020\|EPI_ISL_629796\|20 | ···················· | ···················· | ···················· | ···················· | ···················· | ···················· |
| Q | 4526 hCoV-19/England/QEUH-AD24C5/2020\|EPI_ISL_629797\|20 | ···················· | ···················· | ···················· | ···················· | ···················· | ···················· |
| Q | 4527 hCoV-19/England/QEUH-AD2E4F/2020\|EPI_ISL_629798\|20 | ···················· | ···················· | ···················· | ···················· | ···················· | ···················· |
| Q | 4528 hCoV-19/England/QEUH-AD300C/2020\|EPI_ISL_629799\|20 | ···················· | ···················· | ···················· | ···················· | ···················· | ···················· |
| Q | 4529 hCoV-19/England/ALDP-A66FB6/2020\|EPI_ISL_630676\|20 | ···················· | ···················· | ···················· | ···················· | ···················· | ···················· |
| Q | 4530 hCoV-19/England/ALDP-A6699A/2020\|EPI_ISL_630678\|20 | ···················· | ···················· | ···················· | ···················· | ···················· | ···················· |
| Q | 4531 hCoV-19/England/ALDP-A66AD3/2020\|EPI_ISL_630679\|20 | ···················· | ···················· | ···················· | ···················· | ···················· | ···················· |
| Q | 4532 hCoV-19/England/ALDP-A66D9E/2020\|EPI_ISL_630680\|20 | ···················· | ···················· | ···················· | ···················· | ···················· | ···················· |
| Q | 4533 hCoV-19/England/ALDP-A66C0A/2020\|EPI_ISL_630681\|20 | ···················· | ···················· | ···················· | ···················· | ···················· | ···················· |
| Q | 4534 hCoV-19/England/ALDP-A67850/2020\|EPI_ISL_630682\|20 | ···················· | ···················· | ···················· | ···················· | ···················· | ···················· |
| Q | 4535 hCoV-19/England/ALDP-A66AF1/2020\|EPI_ISL_630683\|20 | ···················· | ···················· | ···················· | ···················· | ···················· | ···················· |
| Q | 4536 hCoV-19/England/ALDP-A68989/2020\|EPI_ISL_630684\|20 | ···················· | ···················· | ···················· | ···················· | ···················· | ···················· |
| Q | 4537 hCoV-19/England/ALDP-A678E7/2020\|EPI_ISL_630686\|20 | ···················· | ···················· | ···················· | ···················· | ···················· | ···················· |
| Q | 4538 hCoV-19/England/ALDP-A66C73/2020\|EPI_ISL_630687\|20 | ···················· | ···················· | ···················· | ···················· | ···················· | ···················· |
| Q | 4539 hCoV-19/England/ALDP-A6797B/2020\|EPI_ISL_630688\|20 | ···················· | ···················· | ···················· | ···················· | ···················· | ···················· |
| Q | 4540 hCoV-19/England/ALDP-A673D7/2020\|EPI_ISL_630689\|20 | ···················· | ···················· | ···················· | ···················· | ···················· | ···················· |
| Q | 4541 hCoV-19/England/ALDP-A6796C/2020\|EPI_ISL_630690\|20 | ···················· | ·A·················· | ···················· | ···················· | ···················· | ···················· |
| Q | 4542 hCoV-19/England/ALDP-A67753/2020\|EPI_ISL_630691\|20 | ···················· | ···················· | ···················· | ···················· | ···················· | ···················· |
| Q | 4543 hCoV-19/England/ALDP-A66FF2/2020\|EPI_ISL_630693\|20 | ···················· | ·A·················· | ···················· | ···················· | ···················· | ···A················ |
| Q | 4544 hCoV-19/England/ALDP-A67A78/2020\|EPI_ISL_630694\|20 | ···················· | ···················· | ···················· | ···················· | ···················· | ···················· |
| Q | 4545 hCoV-19/England/ALDP-A66EE6/2020\|EPI_ISL_630695\|20 | ···················· | ···················· | ···················· | ···················· | ···················· | ···················· |
| Q | 4546 hCoV-19/England/ALDP-A66E13/2020\|EPI_ISL_630696\|20 | ···················· | ···················· | ···················· | ···················· | ···················· | ···················· |
| Q | 4547 hCoV-19/England/ALDP-A67B2A/2020\|EPI_ISL_630697\|20 | ···················· | ···················· | ···················· | ···················· | ···················· | ···················· |
| Q | 4548 hCoV-19/England/ALDP-A67076/2020\|EPI_ISL_630698\|20 | ···················· | ···················· | ···················· | ···················· | ···················· | ···················· |
| Q | 4549 hCoV-19/England/ALDP-A6700D/2020\|EPI_ISL_630699\|20 | ···················· | ···················· | ···················· | ···················· | ···················· | ···················· |
| Q | 4550 hCoV-19/England/ALDP-AA26AA/2020\|EPI_ISL_630713\|20 | ···················· | ···················· | ···················· | ···················· | ···················· | ···················· |
| Q | 4551 hCoV-19/England/ALDP-AA4F70/2020\|EPI_ISL_630714\|20 | ···················· | ···················· | ···················· | ···················· | ···················· | ···················· |
| Q | 4552 hCoV-19/England/ALDP-AA2561/2020\|EPI_ISL_630715\|20 | ···················· | ···················· | ···················· | ···················· | ···················· | ···················· |
| Q | 4553 hCoV-19/England/ALDP-AA2947/2020\|EPI_ISL_630716\|20 | ···················· | ···················· | ···················· | ···················· | ···················· | ···················· |
| Q | 4554 hCoV-19/England/ALDP-AA2A08/2020\|EPI_ISL_630717\|20 | ···················· | ···················· | ···················· | ···················· | ···················· | ···················· |
| Q | 4555 hCoV-19/England/QEUH-AAE644/2020\|EPI_ISL_630718\|20 | ···················· | ···················· | ···················· | ···················· | ···················· | ···················· |
| Q | 4556 hCoV-19/England/QEUH-AAE6AE/2020\|EPI_ISL_630719\|20 | ···················· | ···················· | ···················· | ···················· | ···················· | ···················· |
| Q | 4557 hCoV-19/England/ALDP-AA4CD3/2020\|EPI_ISL_630723\|20 | ···················· | ···················· | ···················· | ···················· | ···················· | ···················· |
| Q | 4558 hCoV-19/England/ALDP-AA50A9/2020\|EPI_ISL_630724\|20 | ···················· | ···················· | ···················· | ···················· | ···················· | ···················· |
| Q | 4559 hCoV-19/England/QEUH-AAE6BD/2020\|EPI_ISL_630725\|20 | ···················· | ···················· | ···················· | ···················· | ···················· | ···················· |
| Q | 4560 hCoV-19/England/ALDP-AA3991/2020\|EPI_ISL_630726\|20 | ···················· | ···················· | ···················· | ···················· | ···················· | ···················· |
| Q | 4561 hCoV-19/England/ALDP-AA30E7/2020\|EPI_ISL_630727\|20 | ···················· | ···················· | ···················· | ···················· | ···················· | ···················· |
| Q | 4562 hCoV-19/England/ALDP-AA321E/2020\|EPI_ISL_630728\|20 | ···················· | ···················· | ···················· | ···················· | ···················· | ···················· |
| Q | 4563 hCoV-19/England/ALDP-AA505E/2020\|EPI_ISL_630729\|20 | ···················· | ···················· | ···················· | ···················· | ···················· | ···················· |
| Q | 4564 hCoV-19/England/ALDP-AA2895/2020\|EPI_ISL_630732\|20 | ···················· | ···················· | ···················· | ···················· | ···················· | ···················· |
| Q | 4565 hCoV-19/England/QEUH-AAE635/2020\|EPI_ISL_630733\|20 | ···················· | ···················· | ···················· | ···················· | ···················· | ···················· |
| Q | 4566 hCoV-19/England/ALDP-AA3FCC/2020\|EPI_ISL_630735\|20 | ···················· | ···················· | ···················· | ···················· | ···················· | ···················· |
| Q | 4567 hCoV-19/England/ALDP-AA4C97/2020\|EPI_ISL_630736\|20 | ···················· | ···················· | ···················· | ···················· | ···················· | ···················· |
| Q | 4568 hCoV-19/England/ALDP-AA4A7F/2020\|EPI_ISL_630737\|20 | ···················· | ···················· | ···················· | ···················· | ···················· | ···················· |
| Q | 4569 hCoV-19/England/ALDP-AA316C/2020\|EPI_ISL_630739\|20 | ···················· | ···················· | ···················· | ···················· | ···················· | ···················· |
| Q | 4570 hCoV-19/England/ALDP-AA1C5E/2020\|EPI_ISL_630740\|20 | ···················· | ···················· | ···················· | ···················· | ···················· | ···················· |
| Q | 4571 hCoV-19/England/ALDP-AA3032/2020\|EPI_ISL_630743\|20 | ···················· | ···················· | ···················· | ···················· | ···················· | ···················· |
| Q | 4572 hCoV-19/England/ALDP-AA3928/2020\|EPI_ISL_630744\|20 | ···················· | ···················· | ···················· | ···················· | ···················· | ···················· |
| Q | 4573 hCoV-19/England/ALDP-AA4D49/2020\|EPI_ISL_630746\|20 | ···················· | ···················· | ···················· | ···················· | ···················· | ···················· |
| Q | 4574 hCoV-19/England/ALDP-AA4CF1/2020\|EPI_ISL_630747\|20 | ···················· | ···················· | ···················· | ···················· | ···················· | ···················· |
| Q | 4575 hCoV-19/England/ALDP-AA4F07/2020\|EPI_ISL_630748\|20 | ···················· | ···················· | ···················· | ···················· | ···················· | ···················· |
| Q | 4576 hCoV-19/England/ALDP-AA1B24/2020\|EPI_ISL_630750\|20 | ···················· | ···················· | ···················· | ···················· | ···················· | ···················· |
| Q | 4577 hCoV-19/England/ALDP-AA1B60/2020\|EPI_ISL_630751\|20 | ···················· | ···················· | ···················· | ···················· | ···················· | ···················· |
| Q | 4578 hCoV-19/England/ALDP-AA315D/2020\|EPI_ISL_630752\|20 | ···················· | ···················· | ···················· | ···················· | ···················· | ···················· |
| Q | 4579 hCoV-19/England/QEUH-AAF0DC/2020\|EPI_ISL_630775\|20 | ···················· | ···················· | ···················· | ···················· | ···················· | ···················· |
| Q | 4580 hCoV-19/England/ALDP-AA2622/2020\|EPI_ISL_630776\|20 | ···················· | ···················· | ···················· | ···················· | ···················· | ···················· |
| Q | 4581 hCoV-19/England/ALDP-AA2938/2020\|EPI_ISL_630777\|20 | ···················· | ···················· | ···················· | ···················· | ···················· | ···················· |
| Q | 4582 hCoV-19/England/QEUH-AAE107/2020\|EPI_ISL_630778\|20 | ···················· | ···················· | ···················· | ···················· | ···················· | ···················· |
| Q | 4583 hCoV-19/England/QEUH-AAE69F/2020\|EPI_ISL_630782\|20 | ···················· | ···················· | ···················· | ···················· | ···················· | ···················· |
| Q | 4584 hCoV-19/England/ALDP-AA2A17/2020\|EPI_ISL_630783\|20 | ···················· | ···················· | ···················· | ···················· | ···················· | ···················· |
| Q | 4585 hCoV-19/England/ALDP-AA4C4C/2020\|EPI_ISL_630784\|20 | ···················· | ···················· | ···················· | ···················· | ···················· | ···················· |
| Q | 4586 hCoV-19/England/ALDP-AA4EEC/2020\|EPI_ISL_630785\|20 | ···················· | ···················· | ···················· | ···················· | ···················· | ···················· |
| Q | 4587 hCoV-19/England/QEUH-AAE0CE/2020\|EPI_ISL_630787\|20 | ···················· | ···················· | ···················· | ···················· | ···················· | ···················· |
| Q | 4588 hCoV-19/England/ALDP-AA4FF8/2020\|EPI_ISL_630788\|20 | ···················· | ···················· | ···················· | ···················· | ···················· | ···················· |
| Q | 4589 hCoV-19/England/ALDP-AA4CC4/2020\|EPI_ISL_630790\|20 | ···················· | ···················· | ···················· | ···················· | ···················· | ···················· |
| Q | 4590 hCoV-19/England/ALDP-AA2631/2020\|EPI_ISL_630792\|20 | ···················· | ···················· | ···················· | ···················· | ···················· | ···················· |
| Q | 4591 hCoV-19/England/ALDP-AA4F43/2020\|EPI_ISL_630793\|20 | ···················· | ···················· | ···················· | ···················· | ···················· | ···················· |
| Q | 4592 hCoV-19/England/ALDP-AA273E/2020\|EPI_ISL_630795\|20 | ···················· | ···················· | ···················· | ···················· | ···················· | ···················· |
| Q | 4593 hCoV-19/England/ALDP-AA38EF/2020\|EPI_ISL_630796\|20 | ···················· | ···················· | ···················· | ···················· | ···················· | ···················· |
| Q | 4594 hCoV-19/Mexico/BCN-ALSR-2537/2020\|EPI_ISL_635540\|2 | ···················· | ·A·················· | ···················· | ···················· | ···················· | ···················· |
| Q | 4595 hCoV-19/India/UT-58319-CSIR-IGIB/2020\|EPI_ISL_6368 | ···················· | ···················· | ···················· | ···················· | ···················· | ···················· |
| Q | 4596 hCoV-19/India/DL-59608-CSIR-IGIB/2020\|EPI_ISL_6368 | ···················· | ·A·················· | ···················· | ···················· | ···················· | ···A················ |
| Q | 4597 hCoV-19/India/UT-65173-CSIR-IGIB/2020\|EPI_ISL_6368 | ···················· | ·A·················· | ···················· | ···················· | ···················· | ···A················ |
| Q | 4598 hCoV-19/India/UT-59267-CSIR-IGIB/2020\|EPI_ISL_6368 | ···················· | ·A·················· | ···················· | ···················· | ···················· | ···A················ |

| | | | | | | | |
|---|---|---|---|---|---|---|---|
| Q | 4599 hCoV-19/India/DL-59108-CSIR-IGIB/2020\|EPI_ISL_6368 | ···················· | ·A·················· | ···················· | ···················· | ···················· | ··A················· |
| Q | 4600 hCoV-19/India/UT-7570807-CSIR-IGIB/2020\|EPI_ISL_63 | ···················· | ·A·················· | ···················· | ···················· | ···················· | ··A················· |
| Q | 4601 hCoV-19/India/UT-59149-CSIR-IGIB/2020\|EPI_ISL_6368 | ···················· | ·A·················· | ···················· | ···················· | ···················· | ··A················· |
| Q | 4602 hCoV-19/Lithuania/MR-LUHS-Eilnr355/2020\|EPI_ISL_63 | ···················· | ···················· | ···················· | ···················· | ···················· | ···················· |
| Q | 4603 hCoV-19/Lithuania/MR-LUHS-Eilnr364/2020\|EPI_ISL_63 | ···················· | ···················· | ···················· | ···················· | ···················· | ···················· |
| Q | 4604 hCoV-19/Lithuania/MR-LUHS-Eilnr365/2020\|EPI_ISL_63 | ···················· | ···················· | ···················· | ···················· | ···················· | ···················· |
| Q | 4605 hCoV-19/Lithuania/MR-LUHS-Eilnr371/2020\|EPI_ISL_63 | ···················· | ···················· | ···················· | ···················· | ···················· | ···················· |
| Q | 4606 hCoV-19/Lithuania/MR-LUHS-Eilnr381/2020\|EPI_ISL_63 | ···················· | ···················· | ···················· | ···················· | ···················· | ···················· |
| Q | 4607 hCoV-19/Lithuania/MR-LUHS-Eilnr401/2020\|EPI_ISL_63 | ···················· | ···················· | ···················· | ···················· | ···················· | ···················· |
| Q | 4608 hCoV-19/Lithuania/MR-LUHS-Eilnr403/2020\|EPI_ISL_63 | ···················· | ···················· | ···················· | ···················· | ···················· | ···················· |
| Q | 4609 hCoV-19/USA/IL-NM-1209/2020\|EPI_ISL_626507\|2020-06 | ···················· | ···················· | ···················· | ···················· | ···················· | ···················· |
| Q | 4610 hCoV-19/USA/IL-NM-1219/2020\|EPI_ISL_626508\|2020-06 | ···················· | ·A·················· | ···················· | ···················· | ···················· | ···················· |
| Q | 4611 hCoV-19/USA/IL-NM-1229/2020\|EPI_ISL_626509\|2020-06 | ···················· | ···················· | ···················· | ···················· | ···················· | ···················· |
| Q | 4612 hCoV-19/USA/IL-NM-1251/2020\|EPI_ISL_626511\|2020-06 | ···················· | ···················· | ···················· | ···················· | ···················· | ···················· |
| Q | 4613 hCoV-19/USA/IL-NM-1371/2020\|EPI_ISL_626514\|2020-04 | ···················· | ···················· | ···················· | ···················· | ···················· | ···················· |
| Q | 4614 hCoV-19/USA/IL-NM-1372/2020\|EPI_ISL_626515\|2020-04 | ···················· | ···················· | ···················· | ···················· | ···················· | ···················· |
| Q | 4615 hCoV-19/USA/IL-NM-1689/2020\|EPI_ISL_626540\|2020-07 | ···················· | ···················· | ···················· | ···················· | ···················· | ···················· |
| Q | 4616 hCoV-19/USA/IL-NM-1703/2020\|EPI_ISL_626542\|2020-07 | ···················· | ··C················· | ···················· | ···················· | ···················· | ···················· |
| Q | 4617 hCoV-19/Chile/AT-CMM-ATAC02/2020\|EPI_ISL_626549\|20 | ···················· | ···················· | ···················· | ···················· | ···················· | ···················· |
| Q | 4618 hCoV-19/Chile/AT-CMM-ATAC03/2020\|EPI_ISL_626550\|20 | ···················· | ···················· | ···················· | ···················· | ···················· | ···················· |
| Q | 4619 hCoV-19/Chile/AT-CMM-ATAC05/2020\|EPI_ISL_626551\|20 | ···················· | ···················· | ···················· | ···················· | ···················· | ···················· |
| Q | 4620 hCoV-19/Chile/AT-CMM-ATAC06/2020\|EPI_ISL_626552\|20 | ···················· | ···················· | ···················· | ···················· | ···················· | ···················· |
| Q | 4621 hCoV-19/Chile/AT-CMM-ATAC07/2020\|EPI_ISL_626553\|20 | ···················· | ···················· | ···················· | ···················· | ···················· | ···················· |
| Q | 4622 hCoV-19/Chile/AT-CMM-ATAC08/2020\|EPI_ISL_626554\|20 | ···················· | ···················· | ···················· | ···················· | ···················· | ···················· |
| Q | 4623 hCoV-19/Chile/AT-CMM-ATAC09/2020\|EPI_ISL_626555\|20 | ···················· | ···················· | ···················· | ···················· | ···················· | ···················· |
| Q | 4624 hCoV-19/Chile/AT-CMM-ATAC10/2020\|EPI_ISL_626556\|20 | ···················· | ···················· | ···················· | ···················· | ···················· | ···················· |
| Q | 4625 hCoV-19/Chile/AT-CMM-ATAC11/2020\|EPI_ISL_626557\|20 | ···················· | ···················· | ···················· | ···················· | ···················· | ···················· |
| Q | 4626 hCoV-19/Chile/AT-CMM-ATAC12/2020\|EPI_ISL_626558\|20 | ···················· | ···················· | ···················· | ···················· | ···················· | ···················· |
| Q | 4627 hCoV-19/Chile/AT-CMM-ATAC13/2020\|EPI_ISL_626559\|20 | ···················· | ···················· | ···················· | ···················· | ···················· | ···················· |
| Q | 4628 hCoV-19/Chile/AT-CMM-ATAC14/2020\|EPI_ISL_626560\|20 | ···················· | ···················· | ···················· | ···················· | ···················· | ···················· |
| Q | 4629 hCoV-19/Chile/AT-CMM-ATAC15/2020\|EPI_ISL_626561\|20 | ···················· | ···················· | ···················· | ···················· | ···················· | ···················· |
| Q | 4630 hCoV-19/Chile/AT-CMM-ATAC16/2020\|EPI_ISL_626562\|20 | ···················· | ···················· | ···················· | ···················· | ···················· | ··A················· |
| Q | 4631 hCoV-19/Chile/AT-CMM-ATAC18/2020\|EPI_ISL_626563\|20 | ···················· | ···················· | ···················· | ···················· | ···················· | ··A················· |
| Q | 4632 hCoV-19/Chile/AT-CMM-ATAC19/2020\|EPI_ISL_626564\|20 | ···················· | ···················· | ···················· | ···················· | ···················· | ···················· |
| Q | 4633 hCoV-19/Chile/AT-CMM-ATAC20/2020\|EPI_ISL_626565\|20 | ···················· | ···················· | ···················· | ···················· | ···················· | ···················· |
| Q | 4634 hCoV-19/Czech Republic/NRL_6846-23/2020\|EPI_ISL_62 | ···················· | ···················· | ···················· | ···················· | ···················· | ···················· |
| Q | 4635 hCoV-19/Czech Republic/NRL_9179-1/2020\|EPI_ISL_626 | ···················· | ···················· | ···················· | ···················· | ···················· | ···················· |
| Q | 4636 hCoV-19/Czech Republic/NRL_9179-4/2020\|EPI_ISL_626 | ···················· | ···················· | ···················· | ···················· | ···················· | ···················· |
| Q | 4637 hCoV-19/Czech Republic/NRL_9179-5/2020\|EPI_ISL_626 | ···················· | ···················· | ···················· | ···················· | ···················· | ···················· |
| Q | 4638 hCoV-19/Czech Republic/NRL_8779/2020\|EPI_ISL_62657 | ···················· | ···················· | ···················· | ···················· | ···················· | ···················· |
| Q | 4639 hCoV-19/Czech Republic/NRL_8798/2020\|EPI_ISL_62657 | ···················· | ···················· | ···················· | ···················· | ···················· | ···················· |
| Q | 4640 hCoV-19/Czech Republic/NRL_8814/2020\|EPI_ISL_62657 | ···················· | ···················· | ···················· | ···················· | ···················· | ···················· |
| Q | 4641 hCoV-19/Czech Republic/NRL_8884/2020\|EPI_ISL_62657 | ···················· | ···················· | ···················· | ···················· | ···················· | ···················· |
| Q | 4642 hCoV-19/Czech Republic/NRL_8923/2020\|EPI_ISL_62657 | ···················· | ···················· | ···················· | ···················· | ···················· | ···················· |
| Q | 4643 hCoV-19/Czech Republic/NRL_8927/2020\|EPI_ISL_62657 | ···················· | ······C············· | ···················· | ···················· | ···················· | ···················· |
| Q | 4644 hCoV-19/Wales/PHWC-4834E7/2020\|EPI_ISL_627827\|2020 | ···················· | ···················· | ···················· | ···················· | ···················· | ···················· |
| Q | 4645 hCoV-19/Wales/PHWC-4834F6/2020\|EPI_ISL_627828\|2020 | ···················· | ···················· | ···················· | ···················· | ···················· | ···················· |
| Q | 4646 hCoV-19/Wales/PHWC-483502/2020\|EPI_ISL_627829\|2020 | ···················· | ···················· | ···················· | ···················· | ···················· | ···················· |
| Q | 4647 hCoV-19/Wales/PHWC-48356C/2020\|EPI_ISL_627832\|2020 | ···················· | ···················· | ···················· | ···················· | ···················· | ···················· |
| Q | 4648 hCoV-19/Wales/PHWC-48357B/2020\|EPI_ISL_627833\|2020 | ···················· | ···················· | ···················· | ···················· | ···················· | ···················· |
| Q | 4649 hCoV-19/Wales/PHWC-48361E/2020\|EPI_ISL_627838\|2020 | ···················· | ···················· | ···················· | ···················· | ···················· | ···················· |
| Q | 4650 hCoV-19/Wales/PHWC-48363C/2020\|EPI_ISL_627839\|2020 | ···················· | ···················· | ···················· | ···················· | ···················· | ···················· |
| Q | 4651 hCoV-19/Wales/PHWC-48364B/2020\|EPI_ISL_627840\|2020 | ···················· | ···················· | ···················· | ···················· | ···················· | ···················· |
| Q | 4653 hCoV-19/Wales/PHWC-483669/2020\|EPI_ISL_627842\|2020 | ···················· | ···················· | ···················· | ···················· | ···················· | ···················· |
| Q | 4656 hCoV-19/Wales/PHWC-483696/2020\|EPI_ISL_627845\|2020 | ···················· | ···················· | ···················· | ···················· | ···················· | ···················· |
| Q | 4657 hCoV-19/Wales/PHWC-4836B4/2020\|EPI_ISL_627846\|2020 | ···················· | ···················· | ···················· | ···················· | ···················· | ···················· |
| Q | 4658 hCoV-19/Wales/PHWC-4836C3/2020\|EPI_ISL_627847\|2020 | ···················· | ···················· | ···················· | ···················· | ···················· | ···················· |
| Q | 4660 hCoV-19/Wales/PHWC-4838EB/2020\|EPI_ISL_627870\|2020 | ···················· | ···················· | ···················· | ···················· | ···················· | ···················· |
| Q | 4661 hCoV-19/Wales/PHWC-4838FA/2020\|EPI_ISL_627871\|2020 | ···················· | ···················· | ···················· | ···················· | ···················· | ···················· |
| Q | 4662 hCoV-19/Wales/PHWC-483906/2020\|EPI_ISL_627872\|2020 | ···················· | ···················· | ···················· | ···················· | ···················· | ···················· |
| Q | 4664 hCoV-19/Wales/PHWC-483933/2020\|EPI_ISL_627874\|2020 | ···················· | ···················· | ···················· | ···················· | ···················· | ···················· |
| Q | 4665 hCoV-19/Wales/PHWC-483951/2020\|EPI_ISL_627875\|2020 | ···················· | ···················· | ···················· | ···················· | ···················· | ···················· |
| Q | 4666 hCoV-19/Wales/PHWC-483960/2020\|EPI_ISL_627876\|2020 | ···················· | ···················· | ···················· | ···················· | ···················· | ···················· |
| Q | 4667 hCoV-19/Wales/PHWC-48397F/2020\|EPI_ISL_627877\|2020 | ···················· | ···················· | ···················· | ···················· | ···················· | ···················· |
| Q | 4669 hCoV-19/Wales/PHWC-48399D/2020\|EPI_ISL_627879\|2020 | ···················· | ···················· | ···················· | ···················· | ···················· | ···················· |
| Q | 4671 hCoV-19/Wales/PHWC-4839CA/2020\|EPI_ISL_627881\|2020 | ···················· | ···················· | ···················· | ···················· | ···················· | ···················· |
| Q | 4672 hCoV-19/Wales/PHWC-4839F7/2020\|EPI_ISL_627882\|2020 | ···················· | ···················· | ···················· | ···················· | ···················· | ···················· |
| Q | 4673 hCoV-19/Wales/PHWC-483A03/2020\|EPI_ISL_627883\|2020 | ···················· | ···················· | ···················· | ···················· | ···················· | ···················· |
| Q | 4674 hCoV-19/Wales/PHWC-483A12/2020\|EPI_ISL_627884\|2020 | ···················· | ···················· | ···················· | ···················· | ···················· | ···················· |
| Q | 4675 hCoV-19/Wales/PHWC-483A21/2020\|EPI_ISL_627885\|2020 | ···················· | ···················· | ···················· | ···················· | ···················· | ···················· |
| Q | 4677 hCoV-19/Wales/PHWC-483A5E/2020\|EPI_ISL_627887\|2020 | ···················· | ···················· | ···················· | ···················· | ···················· | ···················· |
| Q | 4678 hCoV-19/Wales/PHWC-483A6D/2020\|EPI_ISL_627888\|2020 | ···················· | ···················· | ···················· | ···················· | ···················· | ···················· |
| Q | 4679 hCoV-19/Wales/PHWC-483A9A/2020\|EPI_ISL_627889\|2020 | ···················· | ···················· | ···················· | ···················· | ···················· | ···················· |
| Q | 4681 hCoV-19/Wales/PHWC-483AB8/2020\|EPI_ISL_627891\|2020 | ···················· | ···················· | ···················· | ···················· | ···················· | ···················· |
| Q | 4682 hCoV-19/Wales/PHWC-483AC7/2020\|EPI_ISL_627892\|2020 | ···················· | ···················· | ···················· | ···················· | ···················· | ···················· |
| Q | 4683 hCoV-19/Wales/PHWC-483AD6/2020\|EPI_ISL_627893\|2020 | ···················· | ···················· | ···················· | ···················· | ···················· | ···················· |
| Q | 4684 hCoV-19/Wales/PHWC-483AE5/2020\|EPI_ISL_627894\|2020 | ···················· | ···················· | ···················· | ···················· | ···················· | ···················· |
| Q | 4685 hCoV-19/Wales/PHWC-483B00/2020\|EPI_ISL_627895\|2020 | ···················· | ···················· | ···················· | ···················· | ···················· | ···················· |
| Q | 4686 hCoV-19/Wales/PHWC-483B1F/2020\|EPI_ISL_627896\|2020 | ···················· | ···················· | ···················· | ···················· | ···················· | ···················· |
| Q | 4687 hCoV-19/Wales/PHWC-483B3D/2020\|EPI_ISL_627897\|2020 | ···················· | ···················· | ···················· | ···················· | ···················· | ···················· |

Q 4688 hCoV-19/Wales/PHWC-483B4C/2020|EPI_ISL_627898|2020 ····················· ····················· ······················ ····················· ····················· ·····················
Q 4689 hCoV-19/Wales/PHWC-483B6A/2020|EPI_ISL_627899|2020 ····················· ····················· ······················ ····················· ····················· ·····················
Q 4690 hCoV-19/England/MILK-ABBC9D/2020|EPI_ISL_631091|20 ····················· ·A··················· ······················ ····················· ····················· ···A·················
Q 4691 hCoV-19/England/MILK-ABC36A/2020|EPI_ISL_631092|20 ····················· ····················· ······················ ····················· ····················· ·····················
Q 4692 hCoV-19/England/ALDP-AC5E12/2020|EPI_ISL_631093|20 ····················· ····················· ······················ ····················· ····················· ·····················
Q 4693 hCoV-19/England/CAMC-AAF528/2020|EPI_ISL_631094|20 ····················· ····················· ······················ ····················· ····················· ·····················
Q 4694 hCoV-19/England/MILK-ABC2F4/2020|EPI_ISL_631095|20 ····················· ····················· ······················ ····················· ····················· ·····················
Q 4695 hCoV-19/England/ALDP-AC120B/2020|EPI_ISL_631096|20 ····················· ····················· ······················ ····················· ····················· ·····················
Q 4696 hCoV-19/England/MILK-ABC045/2020|EPI_ISL_631098|20 ····················· ····················· ······················ ····················· ······T·············· ·····················
Q 4697 hCoV-19/England/ALDP-AC5DF7/2020|EPI_ISL_631099|20 ····················· ····················· ······················ ····················· ····················· ·····················
Q 4698 hCoV-19/England/QEUH-AD675F/2020|EPI_ISL_634200|20 ····················· ····················· ······················ ····················· ····················· ·····················
Q 4699 hCoV-19/England/QEUH-AD3BDE/2020|EPI_ISL_634203|20 ····················· ····················· ······················ ····················· ····················· ·····················
Q 4700 hCoV-19/England/QEUH-AD2A3C/2020|EPI_ISL_634204|20 ····················· ····················· ······················ ····················· ····················· ·····················
Q 4701 hCoV-19/England/QEUH-AD3B38/2020|EPI_ISL_634205|20 ····················· ····················· ······················ ····················· ····················· ·····················
Q 4702 hCoV-19/England/QEUH-AD0236/2020|EPI_ISL_634206|20 ····················· ····················· ······················ ····················· ····················· ·····················
Q 4703 hCoV-19/England/QEUH-AD3B65/2020|EPI_ISL_634207|20 ····················· ····················· ······················ ····················· ····················· ·····················
Q 4704 hCoV-19/England/QEUH-AD40ED/2020|EPI_ISL_634208|20 ····················· ····················· ······················ ····················· ····················· ·····················
Q 4705 hCoV-19/England/QEUH-AD04D6/2020|EPI_ISL_634209|20 ····················· ····················· ······················ ····················· ····················· ·····················
Q 4706 hCoV-19/England/QEUH-AD491F/2020|EPI_ISL_634210|20 ····················· ····················· ······················ ····················· ····················· ·····················
Q 4707 hCoV-19/England/QEUH-AD669E/2020|EPI_ISL_634211|20 ····················· ····················· ······················ ····················· ····················· ·····················
Q 4708 hCoV-19/England/QEUH-AD37BC/2020|EPI_ISL_634212|20 ····················· ····················· ······················ ····················· ····················· ·····················
Q 4709 hCoV-19/England/QEUH-AD33F4/2020|EPI_ISL_634213|20 ····················· ····················· ······················ ····················· ····················· ·····················
Q 4710 hCoV-19/England/QEUH-AD2902/2020|EPI_ISL_634214|20 ····················· ····················· ······················ ····················· ····················· ·····················
Q 4711 hCoV-19/England/QEUH-AD5082/2020|EPI_ISL_634215|20 ····················· ····················· ······················ ····················· ····················· ·····················
Q 4712 hCoV-19/England/QEUH-AD2BC0/2020|EPI_ISL_634216|20 ····················· ····················· ······················ ····················· ····················· ·····················
Q 4713 hCoV-19/England/QEUH-AD23F5/2020|EPI_ISL_634217|20 ····················· ····················· ······················ ····················· ····················· ·····················
Q 4714 hCoV-19/England/QEUH-AD2BFD/2020|EPI_ISL_634218|20 ····················· ····················· ······················ ····················· ····················· ·····················
Q 4715 hCoV-19/England/QEUH-AD5073/2020|EPI_ISL_634219|20 ····················· ····················· ······················ ····················· ····················· ·····················
Q 4716 hCoV-19/England/QEUH-AD3D14/2020|EPI_ISL_634220|20 ····················· ····················· ······················ ····················· ····················· ·····················
Q 4717 hCoV-19/England/QEUH-AD3C26/2020|EPI_ISL_634222|20 ····················· ····················· ······················ ····················· ····················· ·····················
Q 4718 hCoV-19/England/QEUH-AD1FF2/2020|EPI_ISL_634223|20 ····················· ····················· ······················ ····················· ····················· ·····················
Q 4719 hCoV-19/England/QEUH-AD0D44/2020|EPI_ISL_634226|20 ····················· ····················· ······················ ····················· ····················· ·····················
Q 4720 hCoV-19/England/QEUH-AD247A/2020|EPI_ISL_634227|20 ····················· ····················· ······················ ····················· ····················· ·····················
Q 4721 hCoV-19/England/QEUH-AD495B/2020|EPI_ISL_634228|20 ····················· ····················· ······················ ····················· ····················· ·····················
Q 4722 hCoV-19/England/QEUH-AD5152/2020|EPI_ISL_634229|20 ····················· ····················· ······················ ····················· ····················· ·····················
Q 4723 hCoV-19/England/QEUH-AD48B8/2020|EPI_ISL_634231|20 ····················· ····················· ······················ ····················· ····················· ·····················
Q 4724 hCoV-19/England/QEUH-AD2CCD/2020|EPI_ISL_634232|20 ····················· ····················· ······················ ····················· ····················· ·····················
Q 4725 hCoV-19/England/QEUH-AD2B93/2020|EPI_ISL_634233|20 ····················· ····················· ······················ ····················· ····················· ·····················
Q 4726 hCoV-19/England/QEUH-AD3BCF/2020|EPI_ISL_634234|20 ····················· ····················· ······················ ····················· ····················· ·····················
Q 4727 hCoV-19/England/QEUH-AD485E/2020|EPI_ISL_634235|20 ····················· ····················· ······················ ····················· ····················· ·····················
Q 4728 hCoV-19/England/QEUH-AD3D23/2020|EPI_ISL_634236|20 ····················· ····················· ······················ ····················· ····················· ·····················
Q 4729 hCoV-19/England/QEUH-AD01EE/2020|EPI_ISL_634237|20 ····················· ····················· ······················ ····················· ····················· ·····················
Q 4730 hCoV-19/England/QEUH-AD37F8/2020|EPI_ISL_634239|20 ····················· ····················· ······················ ····················· ····················· ·····················
Q 4731 hCoV-19/Mexico/BCN-ALSR-2491/2020|EPI_ISL_635500|2 ····················· ·A··················· ······················ ····················· ····················· ·····················
Q 4732 hCoV-19/Mexico/BCN-ALSR-2492/2020|EPI_ISL_635501|2 ····················· ·A··················· ······················ ····················· ····················· ·····················
Q 4733 hCoV-19/Mexico/BCN-ALSR-2493/2020|EPI_ISL_635502|2 ····················· ·A··················· ······················ ····················· ····················· ·····················
Q 4734 hCoV-19/Mexico/BCN-ALSR-2495/2020|EPI_ISL_635504|2 ····················· ·A··················· ······················ ····················· ····················· ·····················
Q 4735 hCoV-19/Mexico/BCN-ALSR-2496/2020|EPI_ISL_635505|2 ····················· ·A··················· ······················ ····················· ····················· ·····················
Q 4736 hCoV-19/Mexico/BCN-ALSR-2497/2020|EPI_ISL_635506|2 ····················· ·A··················· ······················ ····················· ····················· ·····················
Q 4737 hCoV-19/Mexico/BCN-ALSR-2499/2020|EPI_ISL_635507|2 ····················· ·A··················· ······················ ····················· ····················· ·····················
Q 4738 hCoV-19/Mexico/BCN-ALSR-2500/2020|EPI_ISL_635508|2 ····················· ·A··················· ······················ ····················· ····················· ·····················
Q 4739 hCoV-19/Mexico/BCN-ALSR-2504/2020|EPI_ISL_635510|2 ····················· ·A··················· ······················ ····················· ····················· ·····················
Q 4740 hCoV-19/Mexico/BCN-ALSR-2506/2020|EPI_ISL_635511|2 ····················· ·A··················· ······················ ····················· ····················· ·····················
Q 4741 hCoV-19/Mexico/BCN-ALSR-2507/2020|EPI_ISL_635512|2 ····················· ·A··················· ······················ ····················· ····················· ·····················
Q 4742 hCoV-19/Mexico/BCN-ALSR-2508/2020|EPI_ISL_635513|2 ····················· ·A··················· ······················ ····················· ····················· ·····················
Q 4743 hCoV-19/Mexico/BCN-ALSR-2510/2020|EPI_ISL_635515|2 ····················· ·A··················· ······················ ····················· ····················· ·····················
Q 4744 hCoV-19/Mexico/BCN-ALSR-2511/2020|EPI_ISL_635516|2 ····················· ·A··················· ······················ ····················· ····················· ·····················
Q 4745 hCoV-19/Mexico/BCN-ALSR-2512/2020|EPI_ISL_635517|2 ····················· ·A··················· ······················ ····················· ····················· ·····················
Q 4746 hCoV-19/Mexico/BCN-ALSR-2513/2020|EPI_ISL_635518|2 ····················· ·A··················· ······················ ····················· ····················· ·····················
Q 4747 hCoV-19/Mexico/BCN-ALSR-2514/2020|EPI_ISL_635519|2 ····················· ·A··················· ······················ ····················· ····················· ·····················
Q 4748 hCoV-19/Mexico/BCN-ALSR-2515/2020|EPI_ISL_635520|2 ····················· ····················· ······················ ····················· ····················· ·····················
Q 4749 hCoV-19/Mexico/BCN-ALSR-2516/2020|EPI_ISL_635521|2 ····················· ·A··················· ······················ ····················· ····················· ·····················
Q 4750 hCoV-19/Mexico/BCN-ALSR-2517/2020|EPI_ISL_635522|2 ····················· ·A··················· ······················ ····················· ····················· ·····················
Q 4751 hCoV-19/Mexico/BCN-ALSR-2519/2020|EPI_ISL_635523|2 ····················· ·A··················· ······················ ····················· ····················· ·····················
Q 4752 hCoV-19/Mexico/BCN-ALSR-2520/2020|EPI_ISL_635524|2 ····················· ·A··················· ······················ ····················· ····················· ·····················
Q 4753 hCoV-19/Mexico/BCN-ALSR-2521/2020|EPI_ISL_635525|2 ····················· ·A··················· ······················ ····················· ····················· ·····················
Q 4754 hCoV-19/Mexico/BCN-ALSR-2522/2020|EPI_ISL_635526|2 ····················· ·A··················· ······················ ····················· ····················· ·····················
Q 4755 hCoV-19/Mexico/BCN-ALSR-2523/2020|EPI_ISL_635527|2 ····················· ····················· ······················ ····················· ····················· ·····················
Q 4756 hCoV-19/Mexico/BCN-ALSR-2524/2020|EPI_ISL_635528|2 ····················· ·A··················· ······················ ····················· ····················· ·····················
Q 4757 hCoV-19/Mexico/BCN-ALSR-2525/2020|EPI_ISL_635529|2 ····················· ·A··················· ······················ ····················· ····················· ·····················
Q 4758 hCoV-19/Mexico/BCN-ALSR-2526/2020|EPI_ISL_635530|2 ····················· ·A··················· ······················ ····················· ····················· ·····················
Q 4759 hCoV-19/Mexico/BCN-ALSR-2528/2020|EPI_ISL_635532|2 ····················· ·A··················· ······················ ····················· ····················· ·····················
Q 4760 hCoV-19/Mexico/BCN-ALSR-2529/2020|EPI_ISL_635533|2 ····················· ·A··················· ······················ ····················· ····················· ·····················
Q 4761 hCoV-19/Mexico/BCN-ALSR-2530/2020|EPI_ISL_635534|2 ····················· ·A··················· ······················ ····················· ····················· ·····················
Q 4762 hCoV-19/Mexico/BCN-ALSR-2531/2020|EPI_ISL_635535|2 ····················· ·A··················· ······················ ····················· ····················· ·····················
Q 4763 hCoV-19/Mexico/BCN-ALSR-2533/2020|EPI_ISL_635536|2 ····················· ····················· ······················ ····················· ····················· ·····················
Q 4764 hCoV-19/Mexico/BCN-ALSR-2534/2020|EPI_ISL_635537|2 ····················· ·A··················· ······················ ····················· ····················· ·····················
Q 4765 hCoV-19/Mexico/BCN-ALSR-2535/2020|EPI_ISL_635538|2 ····················· ·A··················· ······················ ····················· ····················· ·····················
Q 4766 hCoV-19/Mexico/BCN-ALSR-2536/2020|EPI_ISL_635539|2 ····················· ·A··················· ······················ ····················· ····················· ·····················
Q 4767 hCoV-19/Mexico/BCN-ALSR-2538/2020|EPI_ISL_635541|2 ····················· ····················· ······················ ····················· ····················· ·····················

| | | | | | | | |
|---|---|---|---|---|---|---|---|
| Q | 4768 hCoV-19/Mexico/BCN-ALSR-2539/2020\|EPI_ISL_635542\|2 | ···················· | ·A·················· | ····················· | ···················· | ···················· | ···················· |
| Q | 4769 hCoV-19/Mexico/BCN-ALSR-2540/2020\|EPI_ISL_635543\|2 | ···················· | ·A·················· | ····················· | ···················· | ···················· | ···················· |
| Q | 4770 hCoV-19/Mexico/BCN-ALSR-2541/2020\|EPI_ISL_635544\|2 | ···················· | ·A·················· | ····················· | ···················· | ···················· | ···················· |
| Q | 4771 hCoV-19/Mexico/BCN-ALSR-2542/2020\|EPI_ISL_635545\|2 | ···················· | ·A·················· | ····················· | ···················· | ···················· | ···················· |
| Q | 4772 hCoV-19/Mexico/BCN-ALSR-2543/2020\|EPI_ISL_635546\|2 | ···················· | ·A·················· | ····················· | ···················· | ···················· | ···················· |
| Q | 4773 hCoV-19/Mexico/BCN-ALSR-2544/2020\|EPI_ISL_635547\|2 | ···················· | ·A·················· | ····················· | ···················· | ···················· | ···················· |
| Q | 4774 hCoV-19/Mexico/BCN-ALSR-2545/2020\|EPI_ISL_635548\|2 | ···················· | ·A·················· | ····················· | ···················· | ···················· | ···················· |
| Q | 4775 hCoV-19/Mexico/BCN-ALSR-2546/2020\|EPI_ISL_635549\|2 | ···················· | ·A·················· | ····················· | ···················· | ···················· | ···················· |
| Q | 4776 hCoV-19/Mexico/BCN-ALSR-2547/2020\|EPI_ISL_635550\|2 | ···················· | ·A·················· | ····················· | ···················· | ···················· | ···················· |
| Q | 4777 hCoV-19/Mexico/BCN-ALSR-2548/2020\|EPI_ISL_635551\|2 | ···················· | ·A·················· | ····················· | ···················· | ···················· | ···················· |
| Q | 4778 hCoV-19/Mexico/BCN-ALSR-2549/2020\|EPI_ISL_635552\|2 | ···················· | ·A·················· | ····················· | ···················· | ···················· | ···················· |
| Q | 4779 hCoV-19/Mexico/BCN-ALSR-2550/2020\|EPI_ISL_635553\|2 | ···················· | ·A·················· | ····················· | ···················· | ···················· | ···················· |
| Q | 4780 hCoV-19/Mexico/BCN-ALSR-2551/2020\|EPI_ISL_635554\|2 | ···················· | ·A·················· | ····················· | ···················· | ···················· | ···················· |
| Q | 4781 hCoV-19/Mexico/BCN-ALSR-2552/2020\|EPI_ISL_635555\|2 | ···················· | ·A·················· | ····················· | ···················· | ···················· | ···················· |
| Q | 4782 hCoV-19/Mexico/BCN-ALSR-2553/2020\|EPI_ISL_635556\|2 | ···················· | ·A·················· | ····················· | ···················· | ···················· | ···················· |
| Q | 4783 hCoV-19/Mexico/BCN-ALSR-2554/2020\|EPI_ISL_635557\|2 | ···················· | ·A·················· | ····················· | ···················· | ···················· | ···················· |
| Q | 4784 hCoV-19/Mexico/BCN-ALSR-2555/2020\|EPI_ISL_635558\|2 | ···················· | ·A·················· | ····················· | ···················· | ···················· | ···················· |
| Q | 4785 hCoV-19/Mexico/BCN-ALSR-2556/2020\|EPI_ISL_635559\|2 | ···················· | ·A·················· | ····················· | ···················· | ···················· | ···················· |
| Q | 4786 hCoV-19/Mexico/BCN-ALSR-2557/2020\|EPI_ISL_635560\|2 | ···················· | ·A·················· | ····················· | ···················· | ···················· | ···················· |
| Q | 4787 hCoV-19/Mexico/BCN-ALSR-2558/2020\|EPI_ISL_635561\|2 | ···················· | ·A·················· | ···A················· | ···················· | ···················· | ···················· |
| Q | 4788 hCoV-19/Mexico/BCN-ALSR-2559/2020\|EPI_ISL_635562\|2 | ···················· | ···················· | ····················· | ···················· | ···················· | ···················· |
| Q | 4789 hCoV-19/Mexico/BCN-ALSR-2560/2020\|EPI_ISL_635563\|2 | ···················· | ·A·················· | ····················· | ···················· | ···················· | ···················· |
| Q | 4790 hCoV-19/Mexico/BCN-ALSR-2561/2020\|EPI_ISL_635564\|2 | ···················· | ·A·················· | ····················· | ···················· | ···················· | ···················· |
| Q | 4791 hCoV-19/Mexico/BCN-ALSR-2562/2020\|EPI_ISL_635565\|2 | ···················· | ···················· | ····················· | ···················· | ···················· | ···················· |
| Q | 4792 hCoV-19/Mexico/BCN-ALSR-2563/2020\|EPI_ISL_635566\|2 | ···················· | ·A·················· | ····················· | ···················· | ···················· | ···················· |
| Q | 4793 hCoV-19/Mexico/BCN-ALSR-2565/2020\|EPI_ISL_635567\|2 | ···················· | ···················· | ····················· | ···················· | ···················· | ···················· |
| Q | 4794 hCoV-19/Mexico/BCN-ALSR-2566/2020\|EPI_ISL_635568\|2 | ···················· | ·A·················· | ····················· | ···················· | ···················· | ···················· |
| Q | 4795 hCoV-19/Mexico/BCN-ALSR-2568/2020\|EPI_ISL_635569\|2 | ···················· | ·A·················· | ····················· | ···················· | ···················· | ···················· |
| Q | 4796 hCoV-19/India/DL-59113-CSIR-IGIB/2020\|EPI_ISL_6368 | ···················· | ·A·················· | ····················· | ···················· | ···················· | ···A················ |
| Q | 4797 hCoV-19/India/DL-59597-CSIR-IGIB/2020\|EPI_ISL_6368 | ···················· | ·A·················· | ····················· | ···················· | ···················· | ···A················ |
| Q | 4798 hCoV-19/India/DL-65029-CSIR-IGIB/2020\|EPI_ISL_6368 | ···················· | ···················· | ····················· | ···················· | ···················· | ···················· |
| Q | 4799 hCoV-19/India/DL-65025-CSIR-IGIB/2020\|EPI_ISL_6368 | ···················· | ·A··············A··· | ····················· | ···················· | ···················· | ···A················ |
| Q | 4800 hCoV-19/India/DL-65054-CSIR-IGIB/2020\|EPI_ISL_6368 | ···················· | ·A·················· | ····················· | ···················· | ···················· | ···A················ |
| Q | 4801 hCoV-19/India/DL-65092-CSIR-IGIB/2020\|EPI_ISL_6368 | ···················· | ···················· | ····················· | ···················· | ···················· | ···················· |
| Q | 4802 hCoV-19/India/DL-65112-CSIR-IGIB/2020\|EPI_ISL_6368 | ···················· | ·A·················· | ····················· | ···················· | ···················· | ···A················ |
| Q | 4803 hCoV-19/India/UT-65121-CSIR-IGIB/2020\|EPI_ISL_6368 | ···················· | ·A·················· | ····················· | ···················· | ···················· | ···A················ |
| Q | 4804 hCoV-19/India/UT-8570838-CSIR-IGIB/2020\|EPI_ISL_63 | ···················· | ·A·················· | ····················· | ···················· | ···················· | ···A················ |
| Q | 4805 hCoV-19/India/UT-65137-CSIR-IGIB/2020\|EPI_ISL_6368 | ···················· | ·A·················· | ····················· | ···················· | ···················· | ···A················ |
| Q | 4806 hCoV-19/India/DL-NCDC-A1-CSIR-IGIB/2020\|EPI_ISL_63 | ···················· | ···················· | ····················· | ···················· | ···················· | ···················· |
| Q | 4807 hCoV-19/India/UT-65158-CSIR-IGIB/2020\|EPI_ISL_6368 | ···················· | ·A·················· | ····················· | ···················· | ···················· | ···A················ |
| Q | 4808 hCoV-19/India/UT-63116-CSIR-IGIB/2020\|EPI_ISL_6368 | ···················· | ·A·················· | ····················· | ···················· | ···················· | ···A················ |
| Q | 4809 hCoV-19/India/UT-59223-CSIR-IGIB/2020\|EPI_ISL_6368 | ···················· | ·A·················· | ····················· | ···················· | ···················· | ···A················ |
| Q | 4810 hCoV-19/India/UT-59189-CSIR-IGIB/2020\|EPI_ISL_6368 | ···················· | ·A·················· | ····················· | ···················· | ···················· | ···A················ |
| Q | 4811 hCoV-19/Brazil/RJ-01020R1/2020\|EPI_ISL_636834\|2020 | ···················· | ···················· | ····················· | ···················· | ···················· | ···················· |
| Q | 4812 hCoV-19/Brazil/RJ-01020R2/2020\|EPI_ISL_636835\|2020 | ···················· | ···················· | ····················· | ···················· | ···················· | ···················· |
| Q | 4813 hCoV-19/Brazil/RJ-0720R2/2020\|EPI_ISL_636837\|2020- | ···················· | ···················· | ····················· | ···················· | ···················· | ···················· |
| Q | 4814 hCoV-19/Lithuania/MR-LUHS-Eilnr173/2020\|EPI_ISL_63 | ···················· | ···················· | ····················· | ···················· | ···················· | ···················· |
| Q | 4815 hCoV-19/Lithuania/MR-LUHS-Eilnr174/2020\|EPI_ISL_63 | ···················· | ···················· | ····················· | ···················· | ···················· | ···················· |
| Q | 4816 hCoV-19/Lithuania/MR-LUHS-Eilnr189/2020\|EPI_ISL_63 | ···················· | ···················· | ····················· | ···················· | ···················· | ···················· |
| Q | 4817 hCoV-19/Lithuania/MR-LUHS-Eilnr192/2020\|EPI_ISL_63 | ···················· | ···················· | ····················· | ···················· | ···················· | ···················· |
| Q | 4818 hCoV-19/Lithuania/MR-LUHS-Eilnr201/2020\|EPI_ISL_63 | ···················· | ···················· | ····················· | ···················· | ···················· | ···················· |
| Q | 4819 hCoV-19/Lithuania/MR-LUHS-Eilnr203/2020\|EPI_ISL_63 | ···················· | ···················· | ····················· | ···················· | ···················· | ···················· |
| Q | 4820 hCoV-19/Lithuania/MR-LUHS-Eilnr214/2020\|EPI_ISL_63 | ···················· | ···················· | ····················· | ···················· | ···················· | ···················· |
| Q | 4821 hCoV-19/Lithuania/MR-LUHS-Eilnr216/2020\|EPI_ISL_63 | ···················· | ···················· | ····················· | ···················· | ···················· | ···················· |
| Q | 4822 hCoV-19/Lithuania/MR-LUHS-Eilnr222/2020\|EPI_ISL_63 | ···················· | ···················· | ····················· | ···················· | ···················· | ···················· |
| Q | 4823 hCoV-19/Lithuania/MR-LUHS-Eilnr227/2020\|EPI_ISL_63 | ···················· | ···················· | ····················· | ···················· | ···················· | ···················· |
| Q | 4824 hCoV-19/Lithuania/MR-LUHS-Eilnr228/2020\|EPI_ISL_63 | ···················· | ···················· | ····················· | ···················· | ···················· | ···················· |
| Q | 4825 hCoV-19/Lithuania/MR-LUHS-Eilnr230/2020\|EPI_ISL_63 | ···················· | ···················· | ····················· | ···················· | ···················· | ···················· |
| Q | 4826 hCoV-19/Lithuania/MR-LUHS-Eilnr236/2020\|EPI_ISL_63 | ···················· | ···················· | ····················· | ···················· | ···················· | ···················· |
| Q | 4827 hCoV-19/Lithuania/MR-LUHS-Eilnr247/2020\|EPI_ISL_63 | ···················· | ···················· | ····················· | ···················· | ···················· | ···················· |
| Q | 4828 hCoV-19/Lithuania/MR-LUHS-Eilnr248/2020\|EPI_ISL_63 | ···················· | ···················· | ····················· | ···················· | ···················· | ···················· |
| Q | 4829 hCoV-19/Lithuania/MR-LUHS-Eilnr249/2020\|EPI_ISL_63 | ···················· | ···················· | ····················· | ···················· | ···················· | ···················· |
| Q | 4830 hCoV-19/Lithuania/MR-LUHS-Eilnr258/2020\|EPI_ISL_63 | ···················· | ···················· | ····················· | ···················· | ···················· | ···················· |
| Q | 4831 hCoV-19/Lithuania/MR-LUHS-Eilnr262/2020\|EPI_ISL_63 | ···················· | ···················· | ····················· | ···················· | ···················· | ···················· |
| Q | 4832 hCoV-19/Lithuania/MR-LUHS-Eilnr269/2020\|EPI_ISL_63 | ···················· | ···················· | ····················· | ···················· | ···················· | ···················· |
| Q | 4833 hCoV-19/Lithuania/MR-LUHS-Eilnr274/2020\|EPI_ISL_63 | ···················· | ···················· | ····················· | ···················· | ···················· | ···················· |
| Q | 4834 hCoV-19/Lithuania/MR-LUHS-Eilnr282/2020\|EPI_ISL_63 | ···················· | ···················· | ····················· | ···················· | ···················· | ···················· |
| Q | 4835 hCoV-19/Lithuania/MR-LUHS-Eilnr285/2020\|EPI_ISL_63 | ···················· | ···················· | ····················· | ···················· | ···················· | ···················· |
| Q | 4836 hCoV-19/Lithuania/MR-LUHS-Eilnr302/2020\|EPI_ISL_63 | ···················· | ···················· | ····················· | ···················· | ···················· | ···················· |
| Q | 4837 hCoV-19/Lithuania/MR-LUHS-Eilnr314/2020\|EPI_ISL_63 | ···················· | ···················· | ····················· | ···················· | ···················· | ···················· |
| Q | 4838 hCoV-19/Lithuania/MR-LUHS-Eilnr319/2020\|EPI_ISL_63 | ···················· | ···················· | ····················· | ···················· | ···················· | ···················· |
| Q | 4839 hCoV-19/Lithuania/MR-LUHS-Eilnr320/2020\|EPI_ISL_63 | ···················· | ···················· | ····················· | ···················· | ···················· | ···················· |
| Q | 4840 hCoV-19/Lithuania/MR-LUHS-Eilnr334/2020\|EPI_ISL_63 | ···················· | ···················· | ····················· | ···················· | ···················· | ···················· |
| Q | 4841 hCoV-19/Lithuania/MR-LUHS-Eilnr335/2020\|EPI_ISL_63 | ···················· | ···················· | ····················· | ···················· | ···················· | ···················· |
| Q | 4842 hCoV-19/Lithuania/MR-LUHS-Eilnr338/2020\|EPI_ISL_63 | ···················· | ···················· | ····················· | ···················· | ···················· | ···················· |
| Q | 4843 hCoV-19/Lithuania/MR-LUHS-Eilnr339/2020\|EPI_ISL_63 | ···················· | ···················· | ····················· | ···················· | ···················· | ···················· |
| Q | 4844 hCoV-19/Lithuania/MR-LUHS-Eilnr340/2020\|EPI_ISL_63 | ···················· | ···················· | ····················· | ···················· | ···················· | ···················· |
| Q | 4845 hCoV-19/Lithuania/MR-LUHS-Eilnr405/2020\|EPI_ISL_63 | ···················· | ···················· | ····················· | ···················· | ···················· | ···················· |
| Q | 4846 hCoV-19/Lithuania/MR-LUHS-Eilnr407/2020\|EPI_ISL_63 | ···················· | ···················· | ····················· | ···················· | ···················· | ···················· |
| Q | 4847 hCoV-19/Lithuania/MR-LUHS-Eilnr409/2020\|EPI_ISL_63 | ···················· | ···················· | ····················· | ···················· | ···················· | ···················· |

| | | | | | | | |
|---|---|---|---|---|---|---|---|
| Q | 4848 hCoV-19/Lithuania/MR-LUHS-Eilnr414/2020\|EPI_ISL_63 | ···················· | ···················· | ···················· | ···················· | ···················· | ···················· |
| Q | 4849 hCoV-19/Lithuania/MR-LUHS-Eilnr426/2020\|EPI_ISL_63 | ···················· | ···················· | ···················· | ···················· | ···················· | ···················· |
| Q | 4850 hCoV-19/Lithuania/MR-LUHS-Eilnr427/2020\|EPI_ISL_63 | ···················· | ···················· | ···················· | ···················· | ···················· | ···················· |
| Q | 4851 hCoV-19/Lithuania/MR-LUHS-Eilnr431/2020\|EPI_ISL_63 | ···················· | ···················· | ···················· | ···················· | ···················· | ···················· |
| Q | 4852 hCoV-19/Lithuania/MR-LUHS-Eilnr442/2020\|EPI_ISL_63 | ···················· | ···················· | ···················· | ···················· | ···················· | ···················· |
| Q | 4853 hCoV-19/Lithuania/MR-LUHS-Eilnr449/2020\|EPI_ISL_63 | ···················· | ···················· | ···················· | ···················· | ···················· | ···················· |
| Q | 4854 hCoV-19/Lithuania/MR-LUHS-Eilnr450/2020\|EPI_ISL_63 | ···················· | ···················· | ···················· | ···················· | ···················· | ···················· |
| Q | 4855 hCoV-19/Lithuania/MR-LUHS-Eilnr452/2020\|EPI_ISL_63 | ···················· | ···················· | ···················· | ···················· | ···················· | ···················· |
| Q | 4856 hCoV-19/Lithuania/MR-LUHS-Eilnr455/2020\|EPI_ISL_63 | ···················· | ···················· | ···················· | ···················· | ···················· | ···················· |
| Q | 4857 hCoV-19/Lithuania/MR-LUHS-Eilnr456/2020\|EPI_ISL_63 | ···················· | ···················· | ···················· | ···················· | ···················· | ···················· |
| Q | 4858 hCoV-19/Lithuania/MR-LUHS-Eilnr458/2020\|EPI_ISL_63 | ···················· | ···················· | ···················· | ···················· | ···················· | ···················· |
| Q | 4859 hCoV-19/Lithuania/MR-LUHS-Eilnr459/2020\|EPI_ISL_63 | ···················· | ···················· | ···················· | ···················· | ···················· | ···················· |
| Q | 4860 hCoV-19/Lithuania/MR-LUHS-Eilnr460/2020\|EPI_ISL_63 | ···················· | ···················· | ···················· | ···················· | ···················· | ···················· |
| Q | 4861 hCoV-19/Lithuania/MR-LUHS-Eilnr461/2020\|EPI_ISL_63 | ···················· | ···················· | ···················· | ···················· | ···················· | ···················· |
| Q | 4862 hCoV-19/Lithuania/MR-LUHS-Eilnr462/2020\|EPI_ISL_63 | ···················· | ···················· | ···················· | ···················· | ···················· | ···················· |
| Q | 4863 hCoV-19/Lithuania/MR-LUHS-Eilnr468/2020\|EPI_ISL_63 | ···················· | ···················· | ···················· | ···················· | ···················· | ···················· |
| Q | 4864 hCoV-19/Lithuania/MR-LUHS-Eilnr469/2020\|EPI_ISL_63 | ···················· | ···················· | ···················· | ···················· | ···················· | ···················· |
| Q | 4865 hCoV-19/USA/IL-NM-1140/2020\|EPI_ISL_626502\|2020-06 | ···················· | ···················· | ···················· | ···················· | ···················· | ···················· |
| Q | 4866 hCoV-19/USA/IL-NM-1157/2020\|EPI_ISL_626503\|2020-06 | ···················· | ···················· | ···················· | ···················· | ···················· | ···················· |
| Q | 4867 hCoV-19/USA/IL-NM-1190/2020\|EPI_ISL_626505\|2020-06 | ···················· | ···················· | ···················· | ···················· | ···················· | ···················· |
| Q | 4868 hCoV-19/USA/IL-NM-1200/2020\|EPI_ISL_626506\|2020-06 | ···················· | ···················· | ···················· | ···················· | ···················· | ···················· |
| Q | 4869 hCoV-19/USA/IL-NM-1437/2020\|EPI_ISL_626521\|2020-07 | ···················· | ···················· | ···················· | ···················· | ···················· | ···················· |
| Q | 4870 hCoV-19/USA/IL-NM-1442/2020\|EPI_ISL_626522\|2020-07 | ···················· | ···················· | ···················· | ···················· | ···················· | ···················· |
| Q | 4871 hCoV-19/USA/IL-NM-1448/2020\|EPI_ISL_626524\|2020-07 | ···················· | ···················· | ···················· | ···················· | ···················· | ···················· |
| Q | 4872 hCoV-19/USA/IL-NM-1466/2020\|EPI_ISL_626525\|2020-07 | ···················· | ···················· | ···················· | ···················· | ···················· | ···················· |
| Q | 4873 hCoV-19/USA/IL-NM-1482/2020\|EPI_ISL_626526\|2020-07 | ···················· | ···················· | ···················· | ···················· | ···················· | ···················· |
| Q | 4874 hCoV-19/USA/IL-NM-1510/2020\|EPI_ISL_626527\|2020-07 | ············T······· | ···················· | ···················· | ···················· | ···················· | ···················· |
| Q | 4875 hCoV-19/USA/IL-NM-1526/2020\|EPI_ISL_626529\|2020-07 | ···················· | ···················· | ···················· | ···················· | ···················· | ···················· |
| Q | 4876 hCoV-19/USA/IL-NM-1555/2020\|EPI_ISL_626532\|2020-07 | ···················· | ·A·················· | ···················· | ···················· | ···················· | ···A················ |
| Q | 4877 hCoV-19/USA/IL-NM-1643/2020\|EPI_ISL_626536\|2020-07 | ···················· | ···················· | ···················· | ···················· | ···················· | ···················· |
| Q | 4878 hCoV-19/USA/IL-NM-1650/2020\|EPI_ISL_626538\|2020-07 | ···················· | ···················· | ···················· | ···················· | ···················· | ···················· |
| Q | 4879 hCoV-19/USA/IL-NM-1673/2020\|EPI_ISL_626539\|2020-07 | ···················· | ···················· | ···················· | ···················· | ···················· | ···················· |
| Q | 4880 hCoV-19/Czech Republic/NRL_8972-1/2020\|EPI_ISL_626 | ···················· | ···················· | ···················· | ···················· | ···················· | ···················· |
| Q | 4881 hCoV-19/Czech Republic/NRL_8972-2/2020\|EPI_ISL_626 | ···················· | ···················· | ···················· | ···················· | ···················· | ···················· |
| Q | 4882 hCoV-19/Czech Republic/NRL_8972-3/2020\|EPI_ISL_626 | ···················· | ···················· | ···················· | ···················· | ···················· | ···················· |
| Q | 4883 hCoV-19/Czech Republic/NRL_8972-5/2020\|EPI_ISL_626 | ···················· | ···················· | ···················· | ···················· | ···················· | ···················· |
| Q | 4884 hCoV-19/Czech Republic/NRL_8997/2020\|EPI_ISL_62658 | ···················· | ···················· | ···················· | ···················· | ···················· | ···················· |
| Q | 4885 hCoV-19/Czech Republic/NRL_9075/2020\|EPI_ISL_62658 | ···················· | ···················· | ···················· | ···················· | ···················· | ···················· |
| Q | 4886 hCoV-19/Czech Republic/NRL_9329/2020\|EPI_ISL_62658 | ···················· | ···················· | ···················· | ···················· | ···················· | ···················· |
| Q | 4887 hCoV-19/Czech Republic/NRL_9384/2020\|EPI_ISL_62658 | ···················· | ···················· | ···················· | ···················· | ···················· | ···················· |
| Q | 4888 hCoV-19/Czech Republic/NRL_9429/2020\|EPI_ISL_62658 | ···················· | ···················· | ···················· | ···················· | ···················· | ···················· |
| Q | 4889 hCoV-19/Czech Republic/NRL_9448/2020\|EPI_ISL_62658 | ···················· | ···················· | ···················· | ···················· | ···················· | ···················· |
| Q | 4890 hCoV-19/Czech Republic/NRL_9505/2020\|EPI_ISL_62659 | ···················· | ···················· | ···················· | ···················· | ···················· | ···················· |
| Q | 4891 hCoV-19/Czech Republic/NRL_9604/2020\|EPI_ISL_62659 | ···················· | ···················· | ···················· | ···················· | ···················· | ···················· |
| Q | 4892 hCoV-19/Czech Republic/NRL_9630/2020\|EPI_ISL_62659 | ···················· | ···················· | ···················· | ···················· | ···················· | ···················· |
| Q | 4893 hCoV-19/Czech Republic/NRL_9640/2020\|EPI_ISL_62659 | ···················· | ···················· | ···················· | ···················· | ···················· | ···················· |
| Q | 4894 hCoV-19/Czech Republic/NRL_9642/2020\|EPI_ISL_62659 | ···················· | ···················· | ···················· | ···················· | ···················· | ···················· |
| Q | 4895 hCoV-19/Czech Republic/NRL_9654/2020\|EPI_ISL_62659 | ···················· | ···················· | ···················· | ···················· | ···················· | ···················· |
| Q | 4896 hCoV-19/Czech Republic/NRL_9663/2020\|EPI_ISL_62659 | ···················· | ···················· | ···················· | ···················· | ···················· | ···················· |
| Q | 4897 hCoV-19/Czech Republic/NRL_9672/2020\|EPI_ISL_62659 | ···················· | ···················· | ···················· | ···················· | ···················· | ···················· |
| Q | 4898 hCoV-19/Czech Republic/NRL_9673/2020\|EPI_ISL_62659 | ···················· | ···················· | ···················· | ···················· | ···················· | ···················· |
| Q | 4899 hCoV-19/Czech Republic/NRL_9677/2020\|EPI_ISL_62659 | ···················· | ···················· | ···················· | ···················· | ···················· | ···················· |
| Q | 4900 hCoV-19/Wales/PHWC-4832ED/2020\|EPI_ISL_627800\|2020 | ···················· | ···················· | ···················· | ···················· | ···················· | ···················· |
| Q | 4901 hCoV-19/Wales/PHWC-4832FC/2020\|EPI_ISL_627801\|2020 | ···················· | ···················· | ···················· | ···················· | ···················· | ···················· |
| Q | 4902 hCoV-19/Wales/PHWC-483317/2020\|EPI_ISL_627802\|2020 | ···················· | ···················· | ···················· | ···················· | ···················· | ···················· |
| Q | 4903 hCoV-19/Wales/PHWC-483326/2020\|EPI_ISL_627803\|2020 | ···················· | ···················· | ···················· | ···················· | ···················· | ···················· |
| Q | 4904 hCoV-19/Wales/PHWC-483335/2020\|EPI_ISL_627804\|2020 | ···················· | ···················· | ···················· | ···················· | ···················· | ···················· |
| Q | 4905 hCoV-19/Wales/PHWC-483344/2020\|EPI_ISL_627805\|2020 | ···················· | ···················· | ···················· | ···················· | ···················· | ···················· |
| Q | 4906 hCoV-19/Wales/PHWC-483353/2020\|EPI_ISL_627806\|2020 | ···················· | ···················· | ···················· | ···················· | ···················· | ···················· |
| Q | 4907 hCoV-19/Wales/PHWC-483362/2020\|EPI_ISL_627807\|2020 | ···················· | ···················· | ···················· | ···················· | ···················· | ···················· |
| Q | 4908 hCoV-19/Wales/PHWC-483371/2020\|EPI_ISL_627808\|2020 | ···················· | ···················· | ···················· | ···················· | ···················· | ···················· |
| Q | 4909 hCoV-19/Wales/PHWC-483380/2020\|EPI_ISL_627809\|2020 | ···················· | ···················· | ···················· | ···················· | ···················· | ···················· |
| Q | 4910 hCoV-19/Wales/PHWC-48339F/2020\|EPI_ISL_627810\|2020 | ···················· | ···················· | ···················· | ···················· | ···················· | ···················· |
| Q | 4912 hCoV-19/Wales/PHWC-4833BD/2020\|EPI_ISL_627812\|2020 | ···················· | ···················· | ···················· | ···················· | ···················· | ···················· |
| Q | 4913 hCoV-19/Wales/PHWC-4833DB/2020\|EPI_ISL_627813\|2020 | ···················· | ···················· | ···················· | ···················· | ···················· | ···················· |
| Q | 4914 hCoV-19/Wales/PHWC-4833EA/2020\|EPI_ISL_627814\|2020 | ···················· | ···················· | ···················· | ···················· | ···················· | ···················· |
| Q | 4915 hCoV-19/Wales/PHWC-4833F9/2020\|EPI_ISL_627815\|2020 | ···················· | ···················· | ···················· | ···················· | ···················· | ···················· |
| Q | 4916 hCoV-19/Wales/PHWC-483405/2020\|EPI_ISL_627816\|2020 | ···················· | ···················· | ···················· | ···················· | ···················· | ···················· |
| Q | 4917 hCoV-19/Wales/PHWC-483414/2020\|EPI_ISL_627817\|2020 | ···················· | ···················· | ···················· | ···················· | ···················· | ···················· |
| Q | 4918 hCoV-19/Wales/PHWC-483423/2020\|EPI_ISL_627818\|2020 | ···················· | ···················· | ···················· | ···················· | ···················· | ···················· |
| Q | 4919 hCoV-19/Wales/PHWC-483432/2020\|EPI_ISL_627819\|2020 | ···················· | ···················· | ···················· | ···················· | ···················· | ···················· |
| Q | 4920 hCoV-19/Wales/PHWC-483441/2020\|EPI_ISL_627820\|2020 | ···················· | ···················· | ···················· | ···················· | ···················· | ···················· |
| Q | 4921 hCoV-19/Wales/PHWC-48346F/2020\|EPI_ISL_627821\|2020 | ···················· | ···················· | ···················· | ···················· | ···················· | ···················· |
| Q | 4922 hCoV-19/Wales/PHWC-48347E/2020\|EPI_ISL_627822\|2020 | ···················· | ···················· | ···················· | ···················· | ···················· | ···················· |
| Q | 4923 hCoV-19/Wales/PHWC-48349C/2020\|EPI_ISL_627823\|2020 | ···················· | ···················· | ···················· | ···················· | ···················· | ···················· |
| Q | 4925 hCoV-19/Wales/PHWC-4834BA/2020\|EPI_ISL_627825\|2020 | ···················· | ···················· | ···················· | ···················· | ···················· | ···················· |
| Q | 4926 hCoV-19/Wales/PHWC-4834D8/2020\|EPI_ISL_627826\|2020 | ···················· | ···················· | ···················· | ···················· | ···················· | ···················· |
| Q | 4927 hCoV-19/Wales/PHWC-483520/2020\|EPI_ISL_627830\|2020 | ···················· | ···················· | ···················· | ···················· | ···················· | ···················· |
| Q | 4928 hCoV-19/Wales/PHWC-48355D/2020\|EPI_ISL_627831\|2020 | ···················· | ···················· | ···················· | ···················· | ···················· | ···················· |
| Q | 4930 hCoV-19/Wales/PHWC-4835B7/2020\|EPI_ISL_627835\|2020 | ···················· | ···················· | ···················· | ···················· | ···················· | ···················· |

Q 4931 hCoV-19/Wales/PHWC-4835E4/2020|EPI_ISL_627836|2020 ···················· ···················· ···················· ···················· ···················· ····················
Q 4932 hCoV-19/Wales/PHWC-48360F/2020|EPI_ISL_627837|2020 ···················· ···················· ···················· ···················· ···················· ····················
Q 4933 hCoV-19/Wales/PHWC-4836E1/2020|EPI_ISL_627849|2020 ···················· ···················· ···················· ···················· ···················· ····················
Q 4934 hCoV-19/Wales/PHWC-4836F0/2020|EPI_ISL_627850|2020 ···················· ···················· ···················· ···················· ···················· ····················
Q 4935 hCoV-19/Wales/PHWC-48370C/2020|EPI_ISL_627851|2020 ···················· ···················· ···················· ···················· ···················· ····················
Q 4936 hCoV-19/Wales/PHWC-48372A/2020|EPI_ISL_627852|2020 ···················· ···················· ···················· ···················· ···················· ····················
Q 4937 hCoV-19/Wales/PHWC-483766/2020|EPI_ISL_627853|2020 ···················· ···················· ···················· ···················· ···················· ····················
Q 4938 hCoV-19/Wales/PHWC-483775/2020|EPI_ISL_627854|2020 ···················· ···················· ···················· ···················· ···················· ····················
Q 4939 hCoV-19/Wales/PHWC-4837A2/2020|EPI_ISL_627855|2020 ···················· ···················· ···················· ···················· ···················· ····················
Q 4940 hCoV-19/Wales/PHWC-4837B1/2020|EPI_ISL_627856|2020 ···················· ···················· ···················· ···················· ···················· ····················
Q 4941 hCoV-19/Wales/PHWC-4837C0/2020|EPI_ISL_627857|2020 ···················· ·A·······A·········· ···················· ···················· ···················· ····················
Q 4942 hCoV-19/Wales/PHWC-4837DF/2020|EPI_ISL_627858|2020 ···················· ···················· ···················· ···················· ···················· ····················
Q 4944 hCoV-19/Wales/PHWC-483818/2020|EPI_ISL_627860|2020 ···················· ···················· ···················· ···················· ···················· ····················
Q 4945 hCoV-19/Wales/PHWC-483845/2020|EPI_ISL_627861|2020 ···················· ···················· ···················· ···················· ···················· ····················
Q 4946 hCoV-19/Wales/PHWC-483854/2020|EPI_ISL_627862|2020 ···················· ···················· ···················· ···················· ···················· ····················
Q 4948 hCoV-19/Wales/PHWC-483872/2020|EPI_ISL_627864|2020 ···················· ···················· ···················· ···················· ···················· ····················
Q 4950 hCoV-19/Wales/PHWC-4838AF/2020|EPI_ISL_627866|2020 ···················· ···················· ···················· ···················· ···················· ····················
Q 4951 hCoV-19/Wales/PHWC-4838BE/2020|EPI_ISL_627867|2020 ···················· ···················· ···················· ···················· ···················· ····················
Q 4953 hCoV-19/Wales/PHWC-4838DC/2020|EPI_ISL_627869|2020 ···················· ···················· ···················· ···················· ···················· ····················
Q 4954 hCoV-19/England/MILK-ABBDF4/2020|EPI_ISL_631000|20 ···················· ···················· ···················· ···················· ···················· ····················
Q 4955 hCoV-19/England/CAMC-AAF582/2020|EPI_ISL_631001|20 ···················· ···················· ···················· ···················· ···················· ····················
Q 4956 hCoV-19/England/MILK-ABBDB8/2020|EPI_ISL_631002|20 ···················· ···················· ···················· ···················· ······T············· ····················
Q 4957 hCoV-19/England/CAMC-AAF75F/2020|EPI_ISL_631003|20 ···················· ···················· ···················· ···················· ···················· ····················
Q 4958 hCoV-19/England/CAMC-AAF652/2020|EPI_ISL_631005|20 ···················· ···················· ···················· ···················· ···················· ····················
Q 4959 hCoV-19/England/MILK-ABB85D/2020|EPI_ISL_631021|20 ···················· ···················· ···················· ···················· ···················· ····················
Q 4960 hCoV-19/England/MILK-ABC2E5/2020|EPI_ISL_631024|20 ···················· ···················· ···················· ···················· ···················· ····················
Q 4961 hCoV-19/England/CAMC-AAF6E9/2020|EPI_ISL_631025|20 ···················· ···················· ···················· ···················· ···················· ····················
Q 4962 hCoV-19/England/MILK-ABC3D3/2020|EPI_ISL_631026|20 ···················· ···················· ···················· ···················· ······T············· ····················
Q 4963 hCoV-19/England/ALDP-AC5D7F/2020|EPI_ISL_631027|20 ···················· ···················· ···················· ···················· ···················· ····················
Q 4964 hCoV-19/England/ALDP-AC12ED/2020|EPI_ISL_631050|20 ···················· ···················· ···················· ···················· ···················· ····················
Q 4965 hCoV-19/England/MILK-ABBC51/2020|EPI_ISL_631051|20 ···················· ···················· ···················· ···················· ···················· ····················
Q 4966 hCoV-19/England/MILK-ABC027/2020|EPI_ISL_631052|20 ···················· ···················· ···················· ···················· ···················· ····················
Q 4967 hCoV-19/England/CAMC-AAF90E/2020|EPI_ISL_631053|20 ···················· ···················· ···················· ···················· ···················· ····················
Q 4968 hCoV-19/England/CAMC-AAF9E0/2020|EPI_ISL_631055|20 ···················· ···················· ···················· ···················· ···················· ····················
Q 4969 hCoV-19/England/ALDP-AC5DCA/2020|EPI_ISL_631056|20 ···················· ···················· ···················· ···················· ···················· ····················
Q 4970 hCoV-19/England/ALDP-ABD27B/2020|EPI_ISL_631057|20 ···················· ···················· ···················· ···················· ···················· ····················
Q 4971 hCoV-19/England/MILK-ABC2D6/2020|EPI_ISL_631058|20 ···················· ···················· ···················· ···················· ···················· ····················
Q 4972 hCoV-19/England/ALDP-AC1247/2020|EPI_ISL_631059|20 ···················· ···················· ···················· ···················· ···················· ····················
Q 4973 hCoV-19/England/ALDP-AC1256/2020|EPI_ISL_631060|20 ···················· ···················· ···················· ···················· ···················· ····················
Q 4974 hCoV-19/England/MILK-ABC221/2020|EPI_ISL_631061|20 ···················· ···················· ···················· ···················· ···················· ····················
Q 4975 hCoV-19/England/CAMC-AAF643/2020|EPI_ISL_631062|20 ···················· ···················· ···················· ···················· ···················· ····················
Q 4976 hCoV-19/England/CAMC-AAF69E/2020|EPI_ISL_631063|20 ···················· ···················· ···················· ···················· ···················· ····················
Q 4977 hCoV-19/England/MILK-ABC18E/2020|EPI_ISL_631064|20 ···················· ···················· ···················· ···················· ···················· ····················
Q 4978 hCoV-19/England/MILK-ABC467/2020|EPI_ISL_631065|20 ···················· ···················· ···················· ···················· ···················· ····················
Q 4979 hCoV-19/England/MILK-ABBE79/2020|EPI_ISL_631066|20 ···················· ···················· ···················· ···················· ······T············· ····················
Q 4980 hCoV-19/England/MILK-ABBDD6/2020|EPI_ISL_631067|20 ···················· ···················· ···················· ···················· ···················· ····················
Q 4981 hCoV-19/England/CAMC-AAF986/2020|EPI_ISL_631069|20 ···················· ···················· ···················· ···················· ···················· ····················
Q 4982 hCoV-19/England/MILK-ABBD7C/2020|EPI_ISL_631070|20 ···················· ···················· ···················· ···················· ······T············· ····················
Q 4983 hCoV-19/England/MILK-ABBE1F/2020|EPI_ISL_631071|20 ···················· ···················· ···················· ···················· ···················· ····················
Q 4984 hCoV-19/England/MILK-ABC494/2020|EPI_ISL_631072|20 ···················· ···················· ···················· ···················· ···················· ····················
Q 4985 hCoV-19/England/MILK-ABBB27/2020|EPI_ISL_631073|20 ···················· ···················· ···················· ···················· ···················· ····················
Q 4986 hCoV-19/England/ALDP-AC12CF/2020|EPI_ISL_631074|20 ···················· ···················· ···················· ···················· ···················· ····················
Q 4987 hCoV-19/England/MILK-ABC397/2020|EPI_ISL_631075|20 ···················· ···················· ···················· ···················· ···················· ····················
Q 4988 hCoV-19/England/MILK-ABC32E/2020|EPI_ISL_631076|20 ···················· ···················· ···················· ···················· ···················· ····················
Q 4989 hCoV-19/England/MILK-ABBF1C/2020|EPI_ISL_631077|20 ···················· ···················· ···················· ···················· ···················· ····················
Q 4990 hCoV-19/England/CAMC-AAF6F8/2020|EPI_ISL_631078|20 ···················· ···················· ···················· ···················· ···················· ····················
Q 4991 hCoV-19/England/MILK-ABBEC4/2020|EPI_ISL_631079|20 ···················· ···················· ···················· ···················· ···················· ····················
Q 4992 hCoV-19/England/CAMC-AAF519/2020|EPI_ISL_631080|20 ···················· ···················· ···················· ···················· ···················· ····················
Q 4993 hCoV-19/England/MILK-ABC41C/2020|EPI_ISL_631082|20 ···················· ···················· ···················· ···················· ···················· ····················
Q 4994 hCoV-19/England/MILK-ABC43A/2020|EPI_ISL_631083|20 ···················· ···················· ···················· ···················· ···················· ····················
Q 4995 hCoV-19/England/ALDP-AC12DE/2020|EPI_ISL_631084|20 ···················· ···················· ···················· ···················· ···················· ····················
Q 4996 hCoV-19/England/MILK-ABC476/2020|EPI_ISL_631085|20 ···················· ···················· ···················· ···················· ···················· ····················
Q 4997 hCoV-19/England/MILK-ABBC15/2020|EPI_ISL_631087|20 ···················· ···················· ···················· ···················· ···················· ····················
Q 4998 hCoV-19/England/MILK-ABBC7F/2020|EPI_ISL_631088|20 ···················· ·A·················· ···················· ···················· ···················· ··A·················
Q 4999 hCoV-19/England/ALDP-ABD2B7/2020|EPI_ISL_631089|20 ···················· ···················· ···················· ···················· ···················· ····················
Q 5000 hCoV-19/USA/MA-JLL-161N/2020|EPI_ISL_632300|2020 ···················· ···················· ···················· ···················· ···················· ····················
Q 5001 hCoV-19/USA/MA-JLL-162N/2020|EPI_ISL_632301|2020 ···················· ···················· ···················· ···················· ···················· ····················
Q 5002 hCoV-19/USA/MA-JLL-164N/2020|EPI_ISL_632302|2020 ···················· ···················· ···················· ···················· ···················· ····················
Q 5003 hCoV-19/USA/MA-JLL-166N/2020|EPI_ISL_632303|2020 ···················· ···················· ···················· ···················· ···················· ····················
Q 5004 hCoV-19/USA/MA-JLL-17O/2020|EPI_ISL_632304|2020 ···················· ···················· ···················· ···················· ···················· ····················
Q 5005 hCoV-19/USA/MA-JLL-17S1/2020|EPI_ISL_632305|2020 ···················· ···················· ···················· ···················· ···················· ····················
Q 5006 hCoV-19/USA/MA-JLL-18S/2020|EPI_ISL_632306|2020 ···················· ···················· ···················· ···················· ···················· ····················
Q 5007 hCoV-19/USA/MA-JLL-19S/2020|EPI_ISL_632307|2020 ···················· ···················· ···················· ···················· ···················· ····················
Q 5008 hCoV-19/USA/MA-JLL-20S/2020|EPI_ISL_632308|2020 ···················· ···················· ···················· ···················· ···················· ····················
Q 5009 hCoV-19/USA/MA-JLL-22S/2020|EPI_ISL_632309|2020 ···················· ···················· ···················· ···················· ···················· ····················
Q 5010 hCoV-19/Tunisia/55006/2020|EPI_ISL_632310|2020-10- ···················· ···················· ···················· ···················· ···················· ····················
Q 5011 hCoV-19/Germany/NW-UBI-47/2020|EPI_ISL_632311|2020 ···················· ···················· ···················· ···················· ···················· ····················
Q 5012 hCoV-19/Sweden/20-11321/2020|EPI_ISL_632312|2020-0 ···················· ···················· ···················· ···················· ···················· ····················
Q 5013 hCoV-19/Sweden/20-73838/2020|EPI_ISL_632313|2020-0 ···················· ···················· ···················· ···················· ···················· ····················
Q 5014 hCoV-19/Netherlands/ZH-EMC-709/2020|EPI_ISL_632389 ···················· ···················· ···················· ···················· ···················· ····················

| | | | | | | | |
|---|---|---|---|---|---|---|---|
| Q | 5015 hCoV-19/Netherlands/un-EMC-792/2020\|EPI_ISL_632390 | ···················· | ···················· | ···················· | ···················· | ·····T·············· | ···················· |
| Q | 5016 hCoV-19/Netherlands/UT-EMC-101/2020\|EPI_ISL_632391 | ···················· | ···················· | ···················· | ···················· | ···················· | ···················· |
| Q | 5017 hCoV-19/England/QEUH-AE2B27/2020\|EPI_ISL_633600\|20 | ···················· | ···················· | ···················· | ···················· | ···················· | ···················· |
| Q | 5018 hCoV-19/England/QEUH-AE319D/2020\|EPI_ISL_633601\|20 | ···················· | ···················· | ···················· | ···················· | ···················· | ···················· |
| Q | 5019 hCoV-19/England/QEUH-AE324F/2020\|EPI_ISL_633602\|20 | ···················· | ···················· | ···················· | ···················· | ···················· | ···················· |
| Q | 5020 hCoV-19/England/QEUH-AE2B09/2020\|EPI_ISL_633603\|20 | ···················· | ···················· | ···················· | ···················· | ···················· | ···················· |
| Q | 5021 hCoV-19/England/QEUH-AE2DC7/2020\|EPI_ISL_633604\|20 | ···················· | ···················· | ···················· | ···················· | ···················· | ···················· |
| Q | 5022 hCoV-19/England/QEUH-AE25ED/2020\|EPI_ISL_633605\|20 | ···················· | ···················· | ···················· | ···················· | ···················· | ···················· |
| Q | 5023 hCoV-19/England/QEUH-AE2CD9/2020\|EPI_ISL_633606\|20 | ···················· | ···················· | ···················· | ···················· | ···················· | ···················· |
| Q | 5024 hCoV-19/England/QEUH-AE1EF2/2020\|EPI_ISL_633607\|20 | ···················· | ···················· | ···················· | ···················· | ···················· | ···················· |
| Q | 5025 hCoV-19/England/QEUH-AE28D5/2020\|EPI_ISL_633608\|20 | ···················· | ···················· | ···················· | ···················· | ···················· | ···················· |
| Q | 5026 hCoV-19/England/QEUH-AE1E6B/2020\|EPI_ISL_633609\|20 | ···················· | ···················· | ···················· | ···················· | ···················· | ···················· |
| Q | 5027 hCoV-19/England/QEUH-AE2C24/2020\|EPI_ISL_633610\|20 | ···················· | ···················· | ···················· | ···················· | ···················· | ···················· |
| Q | 5028 hCoV-19/England/QEUH-AE2BCD/2020\|EPI_ISL_633611\|20 | ···················· | ···················· | ···················· | ···················· | ···················· | ···················· |
| Q | 5029 hCoV-19/England/QEUH-AE2082/2020\|EPI_ISL_633612\|20 | ···················· | ···················· | ···················· | ···················· | ···················· | ···················· |
| Q | 5030 hCoV-19/England/QEUH-AE24E0/2020\|EPI_ISL_633613\|20 | ···················· | ···················· | ···················· | ···················· | ···················· | ···················· |
| Q | 5031 hCoV-19/England/QEUH-AE2C9D/2020\|EPI_ISL_633614\|20 | ···················· | ···················· | ···················· | ···················· | ···················· | ···················· |
| Q | 5032 hCoV-19/England/QEUH-AE2E4C/2020\|EPI_ISL_633616\|20 | ···················· | ···················· | ···················· | ···················· | ···················· | ···················· |
| Q | 5033 hCoV-19/England/QEUH-AE2820/2020\|EPI_ISL_633617\|20 | ···················· | ···················· | ···················· | ···················· | ···················· | ···················· |
| Q | 5034 hCoV-19/England/QEUH-AE2073/2020\|EPI_ISL_633618\|20 | ···················· | ···················· | ···················· | ···················· | ···················· | ···················· |
| Q | 5035 hCoV-19/England/QEUH-AE2EF1/2020\|EPI_ISL_633619\|20 | ···················· | ···················· | ···················· | ···················· | ···················· | ···················· |
| Q | 5036 hCoV-19/England/QEUH-AE30EB/2020\|EPI_ISL_633620\|20 | ···················· | ···················· | ···················· | ···················· | ···················· | ···················· |
| Q | 5037 hCoV-19/England/QEUH-AE22F5/2020\|EPI_ISL_633621\|20 | ···················· | ···················· | ···················· | ···················· | ···················· | ···················· |
| Q | 5038 hCoV-19/England/QEUH-AE21CB/2020\|EPI_ISL_633622\|20 | ···················· | ···················· | ···················· | ···················· | ···················· | ···················· |
| Q | 5039 hCoV-19/England/QEUH-AE29B4/2020\|EPI_ISL_633623\|20 | ···················· | ···················· | ···················· | ···················· | ···················· | ···················· |
| Q | 5040 hCoV-19/England/QEUH-AE2240/2020\|EPI_ISL_633624\|20 | ···················· | ···················· | ···················· | ···················· | ···················· | ···················· |
| Q | 5041 hCoV-19/England/QEUH-AE30CD/2020\|EPI_ISL_633625\|20 | ···················· | ···················· | ···················· | ···················· | ···················· | ···················· |
| Q | 5042 hCoV-19/England/QEUH-AE2F67/2020\|EPI_ISL_633626\|20 | ···················· | ···················· | ···················· | ···················· | ···················· | ···················· |
| Q | 5043 hCoV-19/England/QEUH-AE2EC4/2020\|EPI_ISL_633627\|20 | ···················· | ···················· | ···················· | ···················· | ···················· | ···················· |
| Q | 5044 hCoV-19/England/QEUH-AE1E3E/2020\|EPI_ISL_633628\|20 | ···················· | ···················· | ···················· | ···················· | ···················· | ···················· |
| Q | 5045 hCoV-19/England/QEUH-AE27D8/2020\|EPI_ISL_633629\|20 | ···················· | ···················· | ···················· | ···················· | ···················· | ···················· |
| Q | 5046 hCoV-19/England/QEUH-AE1B55/2020\|EPI_ISL_633630\|20 | ···················· | ···················· | ···················· | ···················· | ···················· | ···················· |
| Q | 5047 hCoV-19/England/QEUH-AE288A/2020\|EPI_ISL_633631\|20 | ···················· | ···················· | ···················· | ···················· | ···················· | ···················· |
| Q | 5048 hCoV-19/England/QEUH-AE2A1B/2020\|EPI_ISL_633632\|20 | ···················· | ···················· | ···················· | ···················· | ···················· | ···················· |
| Q | 5049 hCoV-19/England/QEUH-AE30DC/2020\|EPI_ISL_633633\|20 | ···················· | ···················· | ···················· | ···················· | ···················· | ···················· |
| Q | 5050 hCoV-19/England/QEUH-AE1E98/2020\|EPI_ISL_633634\|20 | ···················· | ···················· | ···················· | ···················· | ···················· | ···················· |
| Q | 5051 hCoV-19/England/QEUH-AE2398/2020\|EPI_ISL_633635\|20 | ···················· | ···················· | ···················· | ···················· | ···················· | ···················· |
| Q | 5052 hCoV-19/England/QEUH-AE1B91/2020\|EPI_ISL_633636\|20 | ···················· | ···················· | ···················· | ···················· | ···················· | ···················· |
| Q | 5053 hCoV-19/England/QEUH-AE21BC/2020\|EPI_ISL_633637\|20 | ···················· | ···················· | ···················· | ···················· | ···················· | ···················· |
| Q | 5054 hCoV-19/England/QEUH-AE2161/2020\|EPI_ISL_633638\|20 | ···················· | ···················· | ···················· | ···················· | ···················· | ···················· |
| Q | 5055 hCoV-19/England/QEUH-AE27E7/2020\|EPI_ISL_633639\|20 | ···················· | ···················· | ···················· | ···················· | ···················· | ···················· |
| Q | 5056 hCoV-19/England/QEUH-AE2A0C/2020\|EPI_ISL_633640\|20 | ···················· | ···················· | ···················· | ···················· | ···················· | ···················· |
| Q | 5057 hCoV-19/England/QEUH-AE200A/2020\|EPI_ISL_633641\|20 | ···················· | ···················· | ···················· | ···················· | ···················· | ···················· |
| Q | 5058 hCoV-19/England/QEUH-AE2538/2020\|EPI_ISL_633643\|20 | ···················· | ···················· | ···················· | ···················· | ···················· | ···················· |
| Q | 5059 hCoV-19/England/QEUH-AE1AD0/2020\|EPI_ISL_633644\|20 | ···················· | ···················· | ···················· | ···················· | ···················· | ···················· |
| Q | 5060 hCoV-19/England/QEUH-AE2AEE/2020\|EPI_ISL_633645\|20 | ···················· | ···················· | ···················· | ···················· | ···················· | ···················· |
| Q | 5061 hCoV-19/England/QEUH-AE2D12/2020\|EPI_ISL_633647\|20 | ···················· | ···················· | ···················· | ···················· | ···················· | ···················· |
| Q | 5062 hCoV-19/England/QEUH-AE2617/2020\|EPI_ISL_633648\|20 | ···················· | ···················· | ···················· | ···················· | ···················· | ···················· |
| Q | 5063 hCoV-19/England/QEUH-AE1D31/2020\|EPI_ISL_633649\|20 | ···················· | ···················· | ···················· | ···················· | ···················· | ···················· |
| Q | 5064 hCoV-19/England/QEUH-AE2B36/2020\|EPI_ISL_633650\|20 | ···················· | ···················· | ···················· | ···················· | ···················· | ···················· |
| Q | 5065 hCoV-19/England/QEUH-AE2732/2020\|EPI_ISL_633651\|20 | ···················· | ···················· | ···················· | ···················· | ···················· | ···················· |
| Q | 5066 hCoV-19/England/QEUH-AE30BE/2020\|EPI_ISL_633652\|20 | ···················· | ···················· | ···················· | ···················· | ···················· | ···················· |
| Q | 5067 hCoV-19/England/QEUH-AE2C06/2020\|EPI_ISL_633654\|20 | ···················· | ···················· | ···················· | ···················· | ···················· | ···················· |
| Q | 5068 hCoV-19/England/QEUH-AE20DD/2020\|EPI_ISL_633655\|20 | ···················· | ···················· | ···················· | ···················· | ···················· | ···················· |
| Q | 5069 hCoV-19/England/QEUH-AE2FB2/2020\|EPI_ISL_633656\|20 | ···················· | ···················· | ···················· | ···················· | ···················· | ···················· |
| Q | 5070 hCoV-19/England/QEUH-AE227D/2020\|EPI_ISL_633657\|20 | ···················· | ···················· | ···················· | ···················· | ···················· | ···················· |
| Q | 5071 hCoV-19/England/QEUH-AE2E3D/2020\|EPI_ISL_633658\|20 | ···················· | ···················· | ···················· | ···················· | ···················· | ···················· |
| Q | 5072 hCoV-19/England/QEUH-AE1CCB/2020\|EPI_ISL_633659\|20 | ···················· | ···················· | ···················· | ···················· | ···················· | ···················· |
| Q | 5073 hCoV-19/England/QEUH-AE2626/2020\|EPI_ISL_633660\|20 | ···················· | ···················· | ···················· | ···················· | ···················· | ···················· |
| Q | 5074 hCoV-19/England/QEUH-AE3054/2020\|EPI_ISL_633661\|20 | ···················· | ···················· | ···················· | ···················· | ···················· | ···················· |
| Q | 5075 hCoV-19/England/QEUH-AE2DB8/2020\|EPI_ISL_633662\|20 | ···················· | ···················· | ···················· | ···················· | ···················· | ···················· |
| Q | 5076 hCoV-19/England/QEUH-AE3151/2020\|EPI_ISL_633663\|20 | ···················· | ···················· | ···················· | ···················· | ···················· | ···················· |
| Q | 5077 hCoV-19/England/QEUH-AE2C33/2020\|EPI_ISL_633664\|20 | ···················· | ···················· | ···················· | ···················· | ···················· | ···················· |
| Q | 5078 hCoV-19/England/QEUH-AE2222/2020\|EPI_ISL_633665\|20 | ···················· | ···················· | ···················· | ···················· | ···················· | ···················· |
| Q | 5079 hCoV-19/England/QEUH-AE1E10/2020\|EPI_ISL_633666\|20 | ···················· | ···················· | ···················· | ···················· | ···················· | ···················· |
| Q | 5080 hCoV-19/England/QEUH-AE1C8F/2020\|EPI_ISL_633667\|20 | ···················· | ···················· | ···················· | ···················· | ···················· | ···················· |
| Q | 5081 hCoV-19/England/QEUH-AE291E/2020\|EPI_ISL_633680\|20 | ···················· | ···················· | ···················· | ···················· | ···················· | ···················· |
| Q | 5082 hCoV-19/England/QEUH-AE2FFE/2020\|EPI_ISL_633681\|20 | ···················· | ···················· | ···················· | ···················· | ···················· | ···················· |
| Q | 5083 hCoV-19/England/QEUH-AE3142/2020\|EPI_ISL_633684\|20 | ···················· | ···················· | ···················· | ···················· | ···················· | ···················· |
| Q | 5084 hCoV-19/England/QEUH-AE3009/2020\|EPI_ISL_633685\|20 | ···················· | ···················· | ···················· | ···················· | ···················· | ···················· |
| Q | 5085 hCoV-19/England/QEUH-AE27BA/2020\|EPI_ISL_633686\|20 | ···················· | ···················· | ···················· | ···················· | ···················· | ···················· |
| Q | 5086 hCoV-19/England/QEUH-AE2143/2020\|EPI_ISL_633687\|20 | ···················· | ···················· | ···················· | ···················· | ···················· | ···················· |
| Q | 5087 hCoV-19/England/QEUH-AE1E5C/2020\|EPI_ISL_633688\|20 | ···················· | ···················· | ···················· | ···················· | ···················· | ···················· |
| Q | 5088 hCoV-19/England/QEUH-AE1DE6/2020\|EPI_ISL_633689\|20 | ···················· | ···················· | ···················· | ···················· | ···················· | ···················· |
| Q | 5089 hCoV-19/USA/UT-UPHL-201113627/2020\|EPI_ISL_634913\| | ···················· | ···················· | ···················· | ···················· | ···················· | ···················· |
| Q | 5090 hCoV-19/USA/UT-UPHL-201111720/2020\|EPI_ISL_634914\| | ···················· | ···················· | ···················· | ···················· | ···················· | ···················· |
| Q | 5091 hCoV-19/USA/UT-UPHL-201123552/2020\|EPI_ISL_634915\| | ···················· | ···················· | ···················· | ···················· | ···················· | ···················· |
| Q | 5092 hCoV-19/USA/UT-UPHL-201132086/2020\|EPI_ISL_634916\| | ···················· | ···················· | ···················· | ···················· | ···················· | ···················· |
| Q | 5093 hCoV-19/USA/UT-UPHL-201123785/2020\|EPI_ISL_634919\| | ···················· | ···················· | ···················· | ···················· | ···················· | ···················· |
| Q | 5094 hCoV-19/USA/UT-UPHL-201130645/2020\|EPI_ISL_634921\| | ···················· | ···················· | ···················· | ···················· | ···················· | ···················· |

| | | | | | | | |
|---|---|---|---|---|---|---|---|
| Q | 5095 hCoV-19/USA/UT-UPHL-201117228/2020\|EPI_ISL_634922\| | ···················· | ···················· | ····················· | ···················· | ···················· | ···················· |
| Q | 5096 hCoV-19/USA/UT-UPHL-201129040/2020\|EPI_ISL_634924\| | ···················· | ···················· | ····················· | ···················· | ···················· | ···················· |
| Q | 5097 hCoV-19/USA/UT-UPHL-201100957/2020\|EPI_ISL_634925\| | ···················· | ···················· | ····················· | ···················· | ···················· | ···················· |
| Q | 5098 hCoV-19/USA/UT-UPHL-201131519/2020\|EPI_ISL_634926\| | ···················· | ·A·················· | ····················· | ···················· | ···················· | ···················· |
| Q | 5099 hCoV-19/USA/UT-UPHL-201132129/2020\|EPI_ISL_634927\| | ···················· | ···················· | ····················· | ···················· | ···················· | ···················· |
| Q | 5100 hCoV-19/USA/UT-UPHL-201131032/2020\|EPI_ISL_634928\| | ···················· | ···················· | ····················· | ···················· | ···················· | ···················· |
| Q | 5101 hCoV-19/USA/UT-UPHL-201107060/2020\|EPI_ISL_634929\| | ···················· | ·A·················· | ····················· | ···················· | ···················· | ···················· |
| Q | 5102 hCoV-19/USA/UT-UPHL-201122052/2020\|EPI_ISL_634931\| | ···················· | ·A·················· | ····················· | ···················· | ···················· | ···················· |
| Q | 5103 hCoV-19/USA/UT-UPHL-201119552/2020\|EPI_ISL_634932\| | ···················· | ·A·················· | ····················· | ···················· | ···················· | ···················· |
| Q | 5104 hCoV-19/Tunisia/55153/2020\|EPI_ISL_634977\|2020-10- | ···················· | ···················· | ····················· | ···················· | ···················· | ···················· |
| Q | 5105 hCoV-19/South Africa/KRISP-K003990/2020\|EPI_ISL_63 | ···················· | ···················· | ····················· | ···················· | ···················· | ···················· |
| Q | 5107 hCoV-19/South Africa/KRISP-K003993/2020\|EPI_ISL_63 | ···················· | ···················· | ····················· | ···················· | ···················· | ···················· |
| Q | 5108 hCoV-19/South Africa/KRISP-K003995/2020\|EPI_ISL_63 | ···················· | ···················· | ····················· | ···················· | ···················· | ···················· |
| Q | 5109 hCoV-19/South Africa/KRISP-K003996/2020\|EPI_ISL_63 | ···················· | ···················· | ····················· | ···················· | ···················· | ···················· |
| Q | 5110 hCoV-19/South Africa/KRISP-K003997/2020\|EPI_ISL_63 | ···················· | ···················· | ····················· | ···················· | ···················· | ···················· |
| Q | 5111 hCoV-19/South Africa/KRISP-K003999/2020\|EPI_ISL_63 | ···················· | ·A·················· | ····················· | ···················· | ···················· | ···················· |
| Q | 5112 hCoV-19/South Africa/KRISP-K004002/2020\|EPI_ISL_63 | ···················· | ·A·················· | ····················· | ···················· | ···················· | ···················· |
| Q | 5113 hCoV-19/South Africa/KRISP-K004004/2020\|EPI_ISL_63 | ···················· | ···················· | ····················· | ···················· | ···················· | ···················· |
| Q | 5114 hCoV-19/South Africa/KRISP-K004005/2020\|EPI_ISL_63 | ···················· | ···················· | ····················· | ···················· | ···················· | ···················· |
| Q | 5115 hCoV-19/South Africa/KRISP-K004008/2020\|EPI_ISL_63 | ···················· | ···················· | ····················· | ···················· | ···················· | ···················· |
| Q | 5116 hCoV-19/South Africa/KRISP-K004009/2020\|EPI_ISL_63 | ···················· | ···················· | ····················· | ···················· | ···················· | ···················· |
| Q | 5117 hCoV-19/South Africa/KRISP-K004010/2020\|EPI_ISL_63 | ···················· | ···················· | ····················· | ···················· | ···················· | ···················· |
| Q | 5118 hCoV-19/South Africa/KRISP-K004011/2020\|EPI_ISL_63 | ·····G·············· | ···················· | ····················· | ···················· | ···················· | ···················· |
| Q | 5119 hCoV-19/South Africa/KRISP-K004012/2020\|EPI_ISL_63 | ···················· | ···················· | ····················· | ···················· | ···················· | ···················· |
| Q | 5120 hCoV-19/South Africa/KRISP-K004013/2020\|EPI_ISL_63 | ···················· | ···················· | ····················· | ···················· | ···················· | ···················· |
| Q | 5121 hCoV-19/USA/IL-NM-0420/2020\|EPI_ISL_626400\|2020-04 | ···················· | ···················· | ····················· | ···················· | ···················· | ···················· |
| Q | 5122 hCoV-19/USA/IL-NM-0422/2020\|EPI_ISL_626401\|2020-04 | ···················· | ···················· | ····················· | ···················· | ···················· | ···················· |
| Q | 5123 hCoV-19/USA/IL-NM-0423/2020\|EPI_ISL_626402\|2020-04 | ···················· | ···················· | ····················· | ···················· | ···················· | ···················· |
| Q | 5124 hCoV-19/USA/IL-NM-0430/2020\|EPI_ISL_626403\|2020-04 | ···················· | ···················· | ····················· | ···················· | ···················· | ···················· |
| Q | 5125 hCoV-19/USA/IL-NM-0431/2020\|EPI_ISL_626404\|2020-04 | ···················· | ···················· | ····················· | ···················· | ···················· | ···················· |
| Q | 5126 hCoV-19/USA/IL-NM-0432/2020\|EPI_ISL_626405\|2020-04 | ···················· | ···················· | ····················· | ···················· | ···················· | ···················· |
| Q | 5127 hCoV-19/USA/IL-NM-0435/2020\|EPI_ISL_626406\|2020-04 | ···················· | ···················· | ····················· | ···················· | ···················· | ···················· |
| Q | 5128 hCoV-19/USA/IL-NM-0436/2020\|EPI_ISL_626407\|2020-04 | ···················· | ···················· | ····················· | ···················· | ···················· | ···················· |
| Q | 5129 hCoV-19/USA/IL-NM-0437/2020\|EPI_ISL_626408\|2020-04 | ···················· | ···················· | ····················· | ···················· | ···················· | ···················· |
| Q | 5130 hCoV-19/USA/IL-NM-0450/2020\|EPI_ISL_626409\|2020-04 | ···················· | ···················· | ····················· | ···················· | ···················· | ···················· |
| Q | 5131 hCoV-19/USA/IL-NM-0451/2020\|EPI_ISL_626410\|2020-04 | ···················· | ···················· | ····················· | ···················· | ···················· | ···················· |
| Q | 5132 hCoV-19/USA/IL-NM-0467/2020\|EPI_ISL_626415\|2020-04 | ···················· | ···················· | ····················· | ···················· | ···················· | ···················· |
| Q | 5133 hCoV-19/USA/IL-NM-0494/2020\|EPI_ISL_626431\|2020-04 | ···················· | ···················· | ····················· | ···················· | ···················· | ···················· |
| Q | 5134 hCoV-19/USA/IL-NM-0515/2020\|EPI_ISL_626438\|2020-04 | ···················· | ···················· | ····················· | ···················· | ···················· | ···················· |
| Q | 5135 hCoV-19/USA/IL-NM-0517/2020\|EPI_ISL_626439\|2020-04 | ···················· | ···················· | ····················· | ···················· | ···················· | ···················· |
| Q | 5136 hCoV-19/USA/IL-NM-0523/2020\|EPI_ISL_626442\|2020-04 | ···················· | ···················· | ····················· | ···················· | ···················· | ···················· |
| Q | 5137 hCoV-19/USA/IL-NM-0530/2020\|EPI_ISL_626443\|2020-04 | ···················· | ···················· | ····················· | ···················· | ···················· | ···················· |
| Q | 5138 hCoV-19/USA/IL-NM-0534/2020\|EPI_ISL_626445\|2020-04 | ···················· | ···················· | ····················· | ···················· | ···················· | ···················· |
| Q | 5139 hCoV-19/USA/IL-NM-0536/2020\|EPI_ISL_626446\|2020-04 | ···················· | ···················· | ····················· | ···················· | ···················· | ···················· |
| Q | 5140 hCoV-19/USA/IL-NM-0539/2020\|EPI_ISL_626447\|2020-04 | ···················· | ···················· | ····················· | ···················· | ···················· | ···················· |
| Q | 5141 hCoV-19/USA/IL-NM-0540/2020\|EPI_ISL_626448\|2020-04 | ···················· | ···················· | ····················· | ···················· | ···················· | ···················· |
| Q | 5142 hCoV-19/USA/IL-NM-0541/2020\|EPI_ISL_626449\|2020-04 | ···················· | ···················· | ····················· | ···················· | ···················· | ···················· |
| Q | 5143 hCoV-19/USA/IL-NM-0542/2020\|EPI_ISL_626450\|2020-04 | ···················· | ···················· | ····················· | ···················· | ···················· | ···················· |
| Q | 5144 hCoV-19/USA/IL-NM-0544/2020\|EPI_ISL_626451\|2020-04 | ···················· | ···················· | ····················· | ···················· | ···················· | ···················· |
| Q | 5145 hCoV-19/USA/IL-NM-0545/2020\|EPI_ISL_626452\|2020-04 | ···················· | ···················· | ····················· | ···················· | ···················· | ···················· |
| Q | 5146 hCoV-19/USA/IL-NM-0546/2020\|EPI_ISL_626453\|2020-04 | ···················· | ···················· | ····················· | ···················· | ···················· | ···················· |
| Q | 5147 hCoV-19/USA/IL-NM-0588/2020\|EPI_ISL_626457\|2020-05 | ···················· | ···················· | ····················· | ···················· | ···················· | ···················· |
| Q | 5148 hCoV-19/USA/IL-NM-0590/2020\|EPI_ISL_626458\|2020-05 | ···················· | ···················· | ····················· | ···················· | ···················· | ···················· |
| Q | 5149 hCoV-19/USA/IL-NM-0606/2020\|EPI_ISL_626459\|2020-05 | ···················· | ···················· | ····················· | ···················· | ···················· | ···················· |
| Q | 5150 hCoV-19/USA/IL-NM-0629/2020\|EPI_ISL_626460\|2020-05 | ···················· | ···················· | ····················· | ···················· | ···················· | ···················· |
| Q | 5151 hCoV-19/USA/IL-NM-0642/2020\|EPI_ISL_626461\|2020-05 | ···················· | ···················· | ····················· | ···················· | ···················· | ···················· |
| Q | 5152 hCoV-19/USA/IL-NM-0672/2020\|EPI_ISL_626464\|2020-03 | ···················· | ···················· | ····················· | ···················· | ···················· | ···················· |
| Q | 5153 hCoV-19/USA/IL-NM-0673/2020\|EPI_ISL_626465\|2020-03 | ···················· | ···················· | ····················· | ···················· | ···················· | ···················· |
| Q | 5154 hCoV-19/USA/IL-NM-0674/2020\|EPI_ISL_626466\|2020-03 | ···················· | ···················· | ····················· | ···················· | ···················· | ···················· |
| Q | 5155 hCoV-19/USA/IL-NM-0675/2020\|EPI_ISL_626467\|2020-03 | ···················· | ···················· | ····················· | ···················· | ···················· | ···················· |
| Q | 5156 hCoV-19/USA/IL-NM-0676/2020\|EPI_ISL_626468\|2020-03 | ···················· | ···················· | ····················· | ···················· | ···················· | ···················· |
| Q | 5157 hCoV-19/USA/IL-NM-0683/2020\|EPI_ISL_626469\|2020-05 | ···················· | ···················· | ····················· | ···················· | ···················· | ···················· |
| Q | 5158 hCoV-19/USA/IL-NM-0693/2020\|EPI_ISL_626470\|2020-05 | ···················· | ···················· | ····················· | ···················· | ···················· | ···················· |
| Q | 5159 hCoV-19/USA/IL-NM-0730/2020\|EPI_ISL_626472\|2020-05 | ···················· | ···················· | ····················· | ···················· | ···················· | ···················· |
| Q | 5160 hCoV-19/USA/IL-NM-0746/2020\|EPI_ISL_626473\|2020-05 | ···················· | ···················· | ····················· | ···················· | ···················· | ···················· |
| Q | 5161 hCoV-19/USA/IL-NM-0793/2020\|EPI_ISL_626476\|2020-05 | ···················· | ···················· | ····················· | ···················· | ···················· | ···················· |
| Q | 5162 hCoV-19/USA/IL-NM-0816/2020\|EPI_ISL_626478\|2020-05 | ···················· | ··········A········· | ····················· | ···················· | ···················· | ···················· |
| Q | 5163 hCoV-19/USA/IL-NM-0856/2020\|EPI_ISL_626480\|2020-05 | ···················· | ···················· | ····················· | ···················· | ···················· | ···················· |
| Q | 5164 hCoV-19/USA/IL-NM-0937/2020\|EPI_ISL_626485\|2020-05 | ···················· | ···················· | ····················· | ···················· | ···················· | ···················· |
| Q | 5165 hCoV-19/USA/IL-NM-0960/2020\|EPI_ISL_626486\|2020-05 | ···················· | ···················· | ····················· | ···················· | ···················· | ···················· |
| Q | 5166 hCoV-19/USA/IL-NM-0982/2020\|EPI_ISL_626487\|2020-05 | ···················· | ···················· | ····················· | ···················· | ···················· | ···················· |
| Q | 5167 hCoV-19/USA/IL-NM-0997/2020\|EPI_ISL_626488\|2020-05 | ···················· | ···················· | ····················· | ···················· | ···················· | ···················· |
| Q | 5168 hCoV-19/USA/IL-NM-1010/2020\|EPI_ISL_626489\|2020-05 | ···················· | ···················· | ····················· | ···················· | ···················· | ···················· |
| Q | 5169 hCoV-19/USA/IL-NM-1047/2020\|EPI_ISL_626492\|2020-06 | ···················· | ···················· | ····················· | ···················· | ···················· | ···················· |
| Q | 5170 hCoV-19/USA/IL-NM-1064/2020\|EPI_ISL_626493\|2020-06 | ···················· | ···················· | ····················· | ···················· | ···················· | ···················· |
| Q | 5171 hCoV-19/USA/IL-NM-1071/2020\|EPI_ISL_626494\|2020-06 | ···················· | ···················· | ····················· | ···················· | ···················· | ···················· |
| Q | 5172 hCoV-19/USA/IL-NM-1076/2020\|EPI_ISL_626495\|2020-06 | ···················· | ···················· | ····················· | ···················· | ···················· | ···················· |
| Q | 5173 hCoV-19/USA/IL-NM-1092/2020\|EPI_ISL_626497\|2020-06 | ···················· | ···················· | ····················· | ···················· | ···················· | ···················· |
| Q | 5174 hCoV-19/USA/IL-NM-1110/2020\|EPI_ISL_626498\|2020-06 | ···················· | ···················· | ····················· | ···················· | ···················· | ···················· |
| Q | 5175 hCoV-19/England/NOTT-1190A9/2020\|EPI_ISL_627706\|20 | ···················· | ···················· | ····················· | ···················· | ···················· | ···················· |

| | | | | | | | |
|---|---|---|---|---|---|---|---|
| Q | 5176 hCoV-19/England/NOTT-1190C7/2020\|EPI_ISL_627707\|20 | ···················· | ···················· | ···················· | ···················· | ···················· | ···················· |
| Q | 5177 hCoV-19/England/NOTT-11949E/2020\|EPI_ISL_627708\|20 | ···················· | ···················· | ···················· | ···················· | ···················· | ···················· |
| Q | 5178 hCoV-19/England/NOTT-119698/2020\|EPI_ISL_627709\|20 | ···················· | ···················· | ···················· | ···················· | ···················· | ···················· |
| Q | 5179 hCoV-19/England/NOTT-1196A7/2020\|EPI_ISL_627710\|20 | ···················· | ···················· | ···················· | ···················· | ···················· | ···················· |
| Q | 5180 hCoV-19/England/NOTT-1196B6/2020\|EPI_ISL_627711\|20 | ···················· | ···················· | ···················· | ···················· | ···················· | ···················· |
| Q | 5181 hCoV-19/England/NOTT-1196E3/2020\|EPI_ISL_627712\|20 | ···················· | ···················· | ···················· | ···················· | ···················· | ···················· |
| Q | 5182 hCoV-19/England/NOTT-1196F2/2020\|EPI_ISL_627713\|20 | ···················· | ···················· | ···················· | ···················· | ···················· | ···················· |
| Q | 5183 hCoV-19/England/NOTT-11971D/2020\|EPI_ISL_627714\|20 | ···················· | ···················· | ···················· | ···················· | ···················· | ···················· |
| Q | 5184 hCoV-19/England/NOTT-11973B/2020\|EPI_ISL_627715\|20 | ···················· | ···················· | ···················· | ···················· | ···················· | ···················· |
| Q | 5185 hCoV-19/England/NOTT-1197B3/2020\|EPI_ISL_627716\|20 | ···················· | ···················· | ···················· | ···················· | ···················· | ···················· |
| Q | 5186 hCoV-19/England/NOTT-1199BD/2020\|EPI_ISL_627717\|20 | ···················· | ···················· | ···················· | ···················· | ···················· | ···················· |
| Q | 5187 hCoV-19/England/NOTT-1199F9/2020\|EPI_ISL_627718\|20 | ···················· | ···················· | ···················· | ···················· | ···················· | ···················· |
| Q | 5188 hCoV-19/England/NOTT-119A14/2020\|EPI_ISL_627719\|20 | ···················· | ···················· | ···················· | ···················· | ···················· | ···················· |
| Q | 5189 hCoV-19/England/NOTT-119A7E/2020\|EPI_ISL_627720\|20 | ···················· | ···················· | ···················· | ···················· | ···················· | ···················· |
| Q | 5190 hCoV-19/England/NOTT-119A8D/2020\|EPI_ISL_627721\|20 | ···················· | ···················· | ···················· | ···················· | ···················· | ···················· |
| Q | 5191 hCoV-19/England/NOTT-119AAB/2020\|EPI_ISL_627722\|20 | ···················· | ···················· | ···················· | ···················· | ···················· | ···················· |
| Q | 5192 hCoV-19/England/NOTT-119ABA/2020\|EPI_ISL_627723\|20 | ···················· | ···················· | ···················· | ···················· | ···················· | ···················· |
| Q | 5193 hCoV-19/England/NOTT-119AC9/2020\|EPI_ISL_627724\|20 | ···················· | ···················· | ···················· | ···················· | ···················· | ···················· |
| Q | 5194 hCoV-19/England/NOTT-119AD8/2020\|EPI_ISL_627725\|20 | ···················· | ···················· | ···················· | ···················· | ···················· | ···················· |
| Q | 5195 hCoV-19/England/NOTT-119B4E/2020\|EPI_ISL_627726\|20 | ···················· | ···················· | ···················· | ···················· | ···················· | ···················· |
| Q | 5196 hCoV-19/England/NOTT-119BD5/2020\|EPI_ISL_627727\|20 | ···················· | ···················· | ···················· | ···················· | ···················· | ···················· |
| Q | 5197 hCoV-19/England/NOTT-119E81/2020\|EPI_ISL_627728\|20 | ···················· | ···················· | ···················· | ···················· | ···················· | ···················· |
| Q | 5198 hCoV-19/England/NOTT-119EDC/2020\|EPI_ISL_627729\|20 | ···················· | ···················· | ···················· | ···················· | ···················· | ···················· |
| Q | 5199 hCoV-19/Wales/PHWC-16CEAD/2020\|EPI_ISL_627730\|2020 | ···················· | ···················· | ···················· | ···················· | ···················· | ···················· |
| Q | 5200 hCoV-19/Wales/PHWC-16D185/2020\|EPI_ISL_627731\|2020 | ···················· | ···················· | ···················· | ···················· | N··················· | ···················· |
| Q | 5201 hCoV-19/Wales/PHWC-482C4A/2020\|EPI_ISL_627732\|2020 | ···················· | ···················· | ···················· | ···················· | ···················· | ···················· |
| Q | 5202 hCoV-19/Wales/PHWC-482D47/2020\|EPI_ISL_627733\|2020 | ···················· | ···················· | ···················· | ···················· | ···················· | ···················· |
| Q | 5203 hCoV-19/Wales/PHWC-482D56/2020\|EPI_ISL_627734\|2020 | ···················· | ···················· | ···················· | ···················· | ···················· | ···················· |
| Q | 5205 hCoV-19/Wales/PHWC-482D74/2020\|EPI_ISL_627736\|2020 | ···················· | ···················· | ···················· | ···················· | ···················· | ···················· |
| Q | 5206 hCoV-19/Wales/PHWC-482D92/2020\|EPI_ISL_627737\|2020 | ···················· | ···················· | ···················· | ···················· | ···················· | ···················· |
| Q | 5208 hCoV-19/Wales/PHWC-482DB0/2020\|EPI_ISL_627739\|2020 | ···················· | ···················· | ···················· | ···················· | ···················· | ···················· |
| Q | 5209 hCoV-19/Wales/PHWC-482DCF/2020\|EPI_ISL_627740\|2020 | ···················· | ···················· | ···················· | ···················· | ···················· | ···················· |
| Q | 5210 hCoV-19/Wales/PHWC-482DDE/2020\|EPI_ISL_627741\|2020 | ···················· | ···················· | ···················· | ···················· | ···················· | ···················· |
| Q | 5211 hCoV-19/Wales/PHWC-482DED/2020\|EPI_ISL_627742\|2020 | ···················· | ···················· | ···················· | ···················· | ···················· | ···················· |
| Q | 5212 hCoV-19/Wales/PHWC-482DFC/2020\|EPI_ISL_627743\|2020 | ···················· | ···················· | ···················· | ···················· | ···················· | ···················· |
| Q | 5214 hCoV-19/Wales/PHWC-482E17/2020\|EPI_ISL_627745\|2020 | ···················· | ···················· | ···················· | ···················· | ···················· | ···················· |
| Q | 5215 hCoV-19/Wales/PHWC-482E26/2020\|EPI_ISL_627746\|2020 | ···················· | ···················· | ···················· | ···················· | ···················· | ···················· |
| Q | 5221 hCoV-19/Wales/PHWC-482EDB/2020\|EPI_ISL_627752\|2020 | ···················· | ···················· | ···················· | ···················· | ···················· | ···················· |
| Q | 5222 hCoV-19/Wales/PHWC-482EEA/2020\|EPI_ISL_627753\|2020 | ···················· | ···················· | ···················· | ···················· | ···················· | ···················· |
| Q | 5223 hCoV-19/Wales/PHWC-482EF9/2020\|EPI_ISL_627754\|2020 | ········T··········· | ···················· | ···················· | ···················· | ···················· | ···················· |
| Q | 5224 hCoV-19/Wales/PHWC-482F05/2020\|EPI_ISL_627755\|2020 | ···················· | ···················· | ···················· | ···················· | ···················· | ···················· |
| Q | 5225 hCoV-19/Wales/PHWC-482F14/2020\|EPI_ISL_627756\|2020 | ···················· | ···················· | ···················· | ···················· | ···················· | ···················· |
| Q | 5226 hCoV-19/Wales/PHWC-482F23/2020\|EPI_ISL_627757\|2020 | ···················· | ···················· | ···················· | ···················· | ···················· | ···················· |
| Q | 5227 hCoV-19/Wales/PHWC-482F32/2020\|EPI_ISL_627758\|2020 | ···················· | ···················· | ···················· | ···················· | ···················· | ···················· |
| Q | 5228 hCoV-19/Wales/PHWC-482F41/2020\|EPI_ISL_627759\|2020 | ···················· | ···················· | ···················· | ···················· | ···················· | ···················· |
| Q | 5229 hCoV-19/Wales/PHWC-482F50/2020\|EPI_ISL_627760\|2020 | ···················· | ···················· | ···················· | ···················· | ···················· | ···················· |
| Q | 5230 hCoV-19/Wales/PHWC-482F6F/2020\|EPI_ISL_627761\|2020 | ···················· | ···················· | ···················· | ···················· | ···················· | ···················· |
| Q | 5231 hCoV-19/Wales/PHWC-482F7E/2020\|EPI_ISL_627762\|2020 | ···················· | ···················· | ···················· | ···················· | ···················· | ···················· |
| Q | 5232 hCoV-19/Wales/PHWC-482F8D/2020\|EPI_ISL_627763\|2020 | ···················· | ···················· | ···················· | ···················· | ···················· | ···················· |
| Q | 5233 hCoV-19/Wales/PHWC-482F9C/2020\|EPI_ISL_627764\|2020 | ···················· | ···················· | ···················· | ···················· | ···················· | ···················· |
| Q | 5234 hCoV-19/Wales/PHWC-482FBA/2020\|EPI_ISL_627765\|2020 | ···················· | ···················· | ···················· | ···················· | ···················· | ···················· |
| Q | 5235 hCoV-19/Wales/PHWC-482FC9/2020\|EPI_ISL_627766\|2020 | ···················· | ···················· | ···················· | ···················· | ···················· | ···················· |
| Q | 5236 hCoV-19/Wales/PHWC-482FD8/2020\|EPI_ISL_627767\|2020 | ···················· | ·A·················· | ·········T·········· | ···················· | ···················· | ··A················· |
| Q | 5237 hCoV-19/Wales/PHWC-482FF6/2020\|EPI_ISL_627768\|2020 | ···················· | ···················· | ···················· | ···················· | ···················· | ···················· |
| Q | 5238 hCoV-19/Wales/PHWC-48302F/2020\|EPI_ISL_627769\|2020 | ···················· | ···················· | ···················· | ···················· | ···················· | ···················· |
| Q | 5239 hCoV-19/Wales/PHWC-48303E/2020\|EPI_ISL_627770\|2020 | ···················· | ···················· | ···················· | ···················· | ···················· | ···················· |
| Q | 5240 hCoV-19/Wales/PHWC-48304D/2020\|EPI_ISL_627771\|2020 | ···················· | ···················· | ···················· | ···················· | ···················· | ···················· |
| Q | 5241 hCoV-19/Wales/PHWC-48305C/2020\|EPI_ISL_627772\|2020 | ···················· | ···················· | ···················· | ···················· | ···················· | ···················· |
| Q | 5242 hCoV-19/Wales/PHWC-48306B/2020\|EPI_ISL_627773\|2020 | ···················· | ···················· | ···················· | ···················· | ···················· | ···················· |
| Q | 5243 hCoV-19/Wales/PHWC-483089/2020\|EPI_ISL_627774\|2020 | ···················· | ···················· | ···················· | ···················· | ···················· | ···················· |
| Q | 5244 hCoV-19/Wales/PHWC-483098/2020\|EPI_ISL_627775\|2020 | ···················· | ···················· | ···················· | ···················· | ···················· | ···················· |
| Q | 5245 hCoV-19/Wales/PHWC-4830A7/2020\|EPI_ISL_627776\|2020 | ···················· | ···················· | ···················· | ···················· | ···················· | ···················· |
| Q | 5246 hCoV-19/Wales/PHWC-4830B6/2020\|EPI_ISL_627777\|2020 | ···················· | ···················· | ···················· | ···················· | ···················· | ···················· |
| Q | 5247 hCoV-19/Wales/PHWC-4830C5/2020\|EPI_ISL_627778\|2020 | ···················· | ···················· | ···················· | ···················· | ···················· | ···················· |
| Q | 5248 hCoV-19/Wales/PHWC-4830D4/2020\|EPI_ISL_627779\|2020 | ···················· | ···················· | ···················· | ···················· | ···················· | ···················· |
| Q | 5249 hCoV-19/Wales/PHWC-4830E3/2020\|EPI_ISL_627780\|2020 | ···················· | ···················· | ···················· | ···················· | ···················· | ···················· |
| Q | 5251 hCoV-19/Wales/PHWC-48311D/2020\|EPI_ISL_627782\|2020 | ···················· | ···················· | ···················· | ···················· | ···················· | ···················· |
| Q | 5252 hCoV-19/Wales/PHWC-48312C/2020\|EPI_ISL_627783\|2020 | ···················· | ···················· | ···················· | ···················· | ···················· | ···················· |
| Q | 5253 hCoV-19/Wales/PHWC-48314A/2020\|EPI_ISL_627784\|2020 | ···················· | ···················· | ···················· | ···················· | ···················· | ···················· |
| Q | 5254 hCoV-19/Wales/PHWC-483159/2020\|EPI_ISL_627785\|2020 | ···················· | ···················· | ···················· | ···················· | ···················· | ···················· |
| Q | 5255 hCoV-19/Wales/PHWC-483177/2020\|EPI_ISL_627786\|2020 | ···················· | ···················· | ···················· | ···················· | ···················· | ···················· |
| Q | 5256 hCoV-19/Wales/PHWC-483186/2020\|EPI_ISL_627787\|2020 | ···················· | ···················· | ···················· | ···················· | ···················· | ···················· |
| Q | 5257 hCoV-19/Wales/PHWC-4831C2/2020\|EPI_ISL_627788\|2020 | ···················· | ···················· | ···················· | ···················· | ···················· | ···················· |
| Q | 5258 hCoV-19/Wales/PHWC-4831E0/2020\|EPI_ISL_627789\|2020 | ···················· | ···················· | ···················· | ···················· | ···················· | ···················· |
| Q | 5259 hCoV-19/Wales/PHWC-4831FF/2020\|EPI_ISL_627790\|2020 | ···················· | ···················· | ···················· | ···················· | ···················· | ···················· |
| Q | 5260 hCoV-19/Wales/PHWC-48320B/2020\|EPI_ISL_627791\|2020 | ···················· | ···················· | ···················· | ···················· | ···················· | ···················· |
| Q | 5261 hCoV-19/Wales/PHWC-48321A/2020\|EPI_ISL_627792\|2020 | ···················· | ···················· | ···················· | ···················· | ···················· | ···················· |
| Q | 5262 hCoV-19/Wales/PHWC-483238/2020\|EPI_ISL_627793\|2020 | ···················· | ···················· | ···················· | ···················· | ···················· | ···················· |
| Q | 5264 hCoV-19/Wales/PHWC-483265/2020\|EPI_ISL_627795\|2020 | ···················· | ···················· | ···················· | ···················· | ···················· | ···················· |
| Q | 5265 hCoV-19/Wales/PHWC-483283/2020\|EPI_ISL_627796\|2020 | ···················· | ···················· | ···················· | ···················· | ···················· | ···················· |

| | | | | | | | |
|---|---|---|---|---|---|---|---|
| Q | 5266 hCoV-19/Wales/PHWC-4832B0/2020\|EPI_ISL_627797\|2020 | .................... | .................... | .................... | .................... | .................... | .................... |
| Q | 5267 hCoV-19/Wales/PHWC-4832CF/2020\|EPI_ISL_627798\|2020 | .................... | .................... | .................... | .................... | .................... | .................... |
| Q | 5269 hCoV-19/England/MILK-ABC018/2020\|EPI_ISL_631006\|20 | .................... | .................... | .................... | .................... | .................... | .................... |
| Q | 5270 hCoV-19/England/MILK-ABBFB2/2020\|EPI_ISL_631007\|20 | .................... | .................... | .................... | .................... | .................... | .................... |
| Q | 5271 hCoV-19/England/CAMC-AAF722/2020\|EPI_ISL_631008\|20 | .................... | .................... | .................... | .................... | .................... | .................... |
| Q | 5272 hCoV-19/England/MILK-ABBD03/2020\|EPI_ISL_631009\|20 | .................... | .................... | .................... | .................... | .................... | .................... |
| Q | 5273 hCoV-19/England/MILK-ABB83F/2020\|EPI_ISL_631010\|20 | .................... | .................... | .................... | .................... | .................... | .................... |
| Q | 5274 hCoV-19/England/ALDP-AC5D51/2020\|EPI_ISL_631012\|20 | .................... | .................... | .................... | .................... | .................... | .................... |
| Q | 5275 hCoV-19/England/MILK-ABBE00/2020\|EPI_ISL_631013\|20 | .................... | .................... | .................... | .................... | .................... | .................... |
| Q | 5276 hCoV-19/England/ALDP-AC5DE8/2020\|EPI_ISL_631014\|20 | .................... | .................... | .................... | .................... | .................... | .................... |
| Q | 5277 hCoV-19/England/MILK-ABBAC0/2020\|EPI_ISL_631015\|20 | .................... | .................... | .................... | .................... | .................... | .................... |
| Q | 5278 hCoV-19/England/CAMC-AAFA65/2020\|EPI_ISL_631016\|20 | .................... | .................... | .................... | .................... | .................... | .................... |
| Q | 5279 hCoV-19/England/ALDP-AC5D24/2020\|EPI_ISL_631018\|20 | .................... | .................... | .................... | .................... | .................... | .................... |
| Q | 5280 hCoV-19/England/MILK-ABC0CD/2020\|EPI_ISL_631019\|20 | .................... | .................... | .................... | .................... | .................... | .................... |
| Q | 5281 hCoV-19/England/MILK-ABBBFA/2020\|EPI_ISL_631020\|20 | .................... | .................... | .................... | .................... | .................... | .................... |
| Q | 5282 hCoV-19/England/MILK-ABBC33/2020\|EPI_ISL_631022\|20 | .................... | .................... | .................... | .................... | .................... | .................... |
| Q | 5283 hCoV-19/England/CAMC-AAF6AD/2020\|EPI_ISL_631028\|20 | .................... | .................... | .................... | .................... | .................... | .................... |
| Q | 5284 hCoV-19/England/MILK-ABC28B/2020\|EPI_ISL_631029\|20 | .................... | .................... | .................... | .................... | .................... | .................... |
| Q | 5285 hCoV-19/England/MILK-ABBB09/2020\|EPI_ISL_631030\|20 | .................... | .................... | .................... | .................... | .................... | .................... |
| Q | 5286 hCoV-19/England/ALDP-AC1265/2020\|EPI_ISL_631031\|20 | .................... | .................... | .................... | .................... | .................... | .................... |
| Q | 5287 hCoV-19/England/MILK-ABC009/2020\|EPI_ISL_631032\|20 | .................... | .A.................. | .................... | .................... | .................... | ...A................ |
| Q | 5288 hCoV-19/England/ALDP-AC5D33/2020\|EPI_ISL_631033\|20 | .................... | .................... | .................... | .................... | .................... | .................... |
| Q | 5289 hCoV-19/England/MILK-ABC34C/2020\|EPI_ISL_631034\|20 | .................... | .................... | .................... | .................... | .................... | .................... |
| Q | 5290 hCoV-19/England/ALDP-AC5E30/2020\|EPI_ISL_631035\|20 | .................... | .................... | .................... | .................... | .................... | .................... |
| Q | 5291 hCoV-19/England/MILK-ABBBBE/2020\|EPI_ISL_631036\|20 | .................... | .................... | .................... | .................... | .................... | .................... |
| Q | 5292 hCoV-19/England/MILK-ABC072/2020\|EPI_ISL_631037\|20 | .................... | .................... | .................... | .................... | .................... | .................... |
| Q | 5293 hCoV-19/England/CAMC-AAF7D7/2020\|EPI_ISL_631038\|20 | .................... | .................... | .................... | .................... | .................... | .................... |
| Q | 5294 hCoV-19/England/CAMC-AAF801/2020\|EPI_ISL_631039\|20 | .................... | .................... | .................... | .................... | .................... | .................... |
| Q | 5295 hCoV-19/England/MILK-ABB574/2020\|EPI_ISL_631040\|20 | .................... | .................... | .................... | .................... | .................... | .................... |
| Q | 5296 hCoV-19/England/ALDP-AC5D15/2020\|EPI_ISL_631041\|20 | .................... | .................... | .................... | .................... | .................... | .................... |
| Q | 5297 hCoV-19/England/MILK-ABBBEB/2020\|EPI_ISL_631042\|20 | .................... | .................... | .................... | .................... | .................... | .................... |
| Q | 5298 hCoV-19/England/MILK-ABC4FE/2020\|EPI_ISL_631043\|20 | .................... | .................... | .................... | .................... | .................... | .................... |
| Q | 5299 hCoV-19/England/ALDP-AC11E0/2020\|EPI_ISL_631044\|20 | .................... | .................... | .................... | .................... | .................... | .................... |
| Q | 5300 hCoV-19/England/CAMC-AAF8A7/2020\|EPI_ISL_631045\|20 | .................... | .................... | .................... | .................... | .................... | .................... |
| Q | 5301 hCoV-19/England/ALDP-AC5E5E/2020\|EPI_ISL_631046\|20 | .................... | .................... | .................... | .................... | .................... | .................... |
| Q | 5302 hCoV-19/England/MILK-ABC1E8/2020\|EPI_ISL_631047\|20 | .................... | .................... | .................... | .................... | .................... | .................... |
| Q | 5303 hCoV-19/England/MILK-ABC26D/2020\|EPI_ISL_631048\|20 | .................... | .................... | .................... | .................... | ......T............. | .................... |
| Q | 5304 hCoV-19/England/MILK-ABBFFE/2020\|EPI_ISL_631049\|20 | .................... | .................... | .................... | .................... | ......T............. | .................... |
| Q | 5305 hCoV-19/England/QEUH-AE2301/2020\|EPI_ISL_633514\|20 | .................... | .................... | .................... | .................... | .................... | .................... |
| Q | 5306 hCoV-19/England/QEUH-AE1B28/2020\|EPI_ISL_633516\|20 | .................... | .................... | .................... | .................... | .................... | .................... |
| Q | 5307 hCoV-19/England/QEUH-AE1EA7/2020\|EPI_ISL_633517\|20 | .................... | .................... | .................... | .................... | .................... | .................... |
| Q | 5308 hCoV-19/England/QEUH-AE3106/2020\|EPI_ISL_633518\|20 | .................... | .................... | .................... | .................... | .................... | .................... |
| Q | 5309 hCoV-19/England/QEUH-AE1CE9/2020\|EPI_ISL_633519\|20 | .................... | .................... | .................... | .................... | ......T............. | .................... |
| Q | 5310 hCoV-19/England/QEUH-AE29E1/2020\|EPI_ISL_633520\|20 | .................... | .................... | .................... | .................... | .................... | .................... |
| Q | 5311 hCoV-19/England/QEUH-AE2556/2020\|EPI_ISL_633521\|20 | .................... | .................... | .................... | .................... | .................... | .................... |
| Q | 5312 hCoV-19/England/QEUH-AE1BEC/2020\|EPI_ISL_633522\|20 | .................... | .................... | .................... | .................... | .................... | .................... |
| Q | 5313 hCoV-19/England/QEUH-AE30AF/2020\|EPI_ISL_633523\|20 | .................... | .................... | .................... | .................... | .................... | .................... |
| Q | 5314 hCoV-19/England/QEUH-AE3090/2020\|EPI_ISL_633524\|20 | .................... | .................... | .................... | .................... | .................... | .................... |
| Q | 5315 hCoV-19/England/QEUH-AE2477/2020\|EPI_ISL_633668\|20 | .................... | .................... | .................... | .................... | .................... | .................... |
| Q | 5316 hCoV-19/England/QEUH-AE2DE5/2020\|EPI_ISL_633669\|20 | .................... | .................... | .................... | .................... | .................... | .................... |
| Q | 5317 hCoV-19/England/QEUH-AE1A94/2020\|EPI_ISL_633670\|20 | .................... | .................... | .................... | .................... | .................... | .................... |
| Q | 5318 hCoV-19/England/QEUH-AE2714/2020\|EPI_ISL_633671\|20 | .................... | .................... | .................... | .................... | .................... | .................... |
| Q | 5319 hCoV-19/England/QEUH-AE2969/2020\|EPI_ISL_633672\|20 | .................... | .................... | .................... | .................... | .................... | .................... |
| Q | 5320 hCoV-19/England/QEUH-AE2AFD/2020\|EPI_ISL_633673\|20 | .................... | .................... | .................... | .................... | .................... | .................... |
| Q | 5321 hCoV-19/England/QEUH-AE2F1C/2020\|EPI_ISL_633674\|20 | .................... | .................... | .................... | .................... | .................... | .................... |
| Q | 5322 hCoV-19/England/QEUH-AE2B45/2020\|EPI_ISL_633675\|20 | .................... | .................... | .................... | .................... | .................... | .................... |
| Q | 5323 hCoV-19/England/QEUH-AE1B46/2020\|EPI_ISL_633676\|20 | .................... | .................... | .................... | .................... | .................... | .................... |
| Q | 5324 hCoV-19/England/QEUH-AE2D6D/2020\|EPI_ISL_633677\|20 | .................... | .................... | .................... | .................... | .................... | .................... |
| Q | 5325 hCoV-19/England/QEUH-AE2E2E/2020\|EPI_ISL_633678\|20 | .................... | .................... | .................... | .................... | .................... | .................... |
| Q | 5326 hCoV-19/England/QEUH-AE2987/2020\|EPI_ISL_633679\|20 | .................... | .................... | .................... | .................... | .................... | .................... |
| Q | 5327 hCoV-19/England/QEUH-AE23C5/2020\|EPI_ISL_633682\|20 | .................... | .................... | .................... | .................... | .................... | .................... |
| Q | 5328 hCoV-19/England/QEUH-AE226E/2020\|EPI_ISL_633683\|20 | .................... | .................... | .................... | .................... | .................... | .................... |
| Q | 5329 hCoV-19/England/QEUH-AE2B72/2020\|EPI_ISL_633690\|20 | .................... | .................... | .................... | .................... | .................... | .................... |
| Q | 5330 hCoV-19/England/QEUH-AE2644/2020\|EPI_ISL_633691\|20 | .................... | .................... | .................... | .................... | .................... | .................... |
| Q | 5331 hCoV-19/England/QEUH-AE25FC/2020\|EPI_ISL_633692\|20 | .................... | .................... | .................... | .................... | .................... | .................... |
| Q | 5332 hCoV-19/England/QEUH-AD1BC1/2020\|EPI_ISL_633694\|20 | .................... | .................... | .................... | .................... | .................... | .................... |
| Q | 5333 hCoV-19/England/QEUH-AD07A0/2020\|EPI_ISL_633695\|20 | .................... | .................... | .................... | .................... | .................... | .................... |
| Q | 5334 hCoV-19/England/QEUH-AD1587/2020\|EPI_ISL_633698\|20 | .................... | .................... | .................... | .................... | .................... | .................... |
| Q | 5335 hCoV-19/England/QEUH-AD15A5/2020\|EPI_ISL_633699\|20 | .................... | .................... | .................... | .................... | .................... | .................... |
| Q | 5336 hCoV-19/USA/MN-MDH-1972/2020\|EPI_ISL_634846\|2020-1 | .................... | .............A...... | .................... | .................... | .................... | .................... |
| Q | 5337 hCoV-19/USA/MN-MDH-1973/2020\|EPI_ISL_634847\|2020-1 | .................... | .............A...... | .................... | .................... | .................... | .................... |
| Q | 5338 hCoV-19/USA/MN-MDH-1974/2020\|EPI_ISL_634848\|2020-1 | .................... | .............A...... | .................... | .................... | .................... | .................... |
| Q | 5339 hCoV-19/USA/MN-MDH-1975/2020\|EPI_ISL_634849\|2020-1 | .................... | .............A...... | .................... | .................... | .................... | .................... |
| Q | 5340 hCoV-19/USA/MN-MDH-1976/2020\|EPI_ISL_634850\|2020-1 | .................... | .............A...... | .................... | .................... | .................... | .................... |
| Q | 5341 hCoV-19/USA/MN-MDH-1977/2020\|EPI_ISL_634851\|2020-1 | .................... | .............A...... | .................... | .................... | .................... | .................... |
| Q | 5342 hCoV-19/USA/MN-MDH-1978/2020\|EPI_ISL_634852\|2020-1 | .................... | .............A...... | .................... | .................... | .................... | .................... |
| Q | 5343 hCoV-19/USA/MN-MDH-1979/2020\|EPI_ISL_634853\|2020-1 | .................... | .............A...... | .................... | .................... | .................... | .................... |
| Q | 5344 hCoV-19/USA/MN-MDH-1980/2020\|EPI_ISL_634854\|2020-1 | .................... | .................... | .................... | .................... | .................... | .................... |
| Q | 5345 hCoV-19/USA/MN-MDH-1981/2020\|EPI_ISL_634855\|2020-1 | .................... | .................... | .................... | .................... | .................... | .................... |
| Q | 5346 hCoV-19/USA/UT-UPHL-201130637/2020\|EPI_ISL_634900\| | .................... | .................... | .................... | .................... | .................... | .................... |

Q 5347 hCoV-19/USA/UT-UPHL-201113557/2020|EPI_ISL_634901| ···················· ···················· ···················· ···················· ···················· ····················
Q 5348 hCoV-19/USA/UT-UPHL-201114453/2020|EPI_ISL_634902| ···················· ···················· ···················· ···················· ···················· ····················
Q 5349 hCoV-19/USA/UT-UPHL-201108842/2020|EPI_ISL_634903| ···················· ···················· ···················· ···················· ···················· ····················
Q 5350 hCoV-19/USA/UT-UPHL-201119439/2020|EPI_ISL_634904| ···················· ···················· ···················· ···················· ···················· ····················
Q 5351 hCoV-19/USA/UT-UPHL-201106608/2020|EPI_ISL_634905| ···················· ···················· ···················· ···················· ···················· ····················
Q 5352 hCoV-19/USA/UT-UPHL-201131031/2020|EPI_ISL_634906| ···················· ···················· ···················· ···················· ···················· ····················
Q 5353 hCoV-19/USA/UT-UPHL-201117437/2020|EPI_ISL_634907| ···················· ···················· ···················· ···················· ···················· ····················
Q 5354 hCoV-19/USA/UT-UPHL-201131260/2020|EPI_ISL_634908| ···················· ···················· ···················· ···················· ···················· ····················
Q 5355 hCoV-19/USA/UT-UPHL-201113837/2020|EPI_ISL_634909| ···················· ···················· ···················· ···················· ···················· ····················
Q 5356 hCoV-19/South Africa/KRISP-K004014/2020|EPI_ISL_63 ···················· ·A·················· ···················· ···················· ···················· ····················
Q 5357 hCoV-19/Romania/BUCHAREST-IMT-1/2020|EPI_ISL_63973 ···················· ···················· ···················· ···················· ···················· ····················
Q 5358 hCoV-19/Japan/TKYT22342/2020|EPI_ISL_649126|2020-0 ···················· ···················· ···················· ···················· ···················· ····················
Q 5359 hCoV-19/Japan/TKYT22355/2020|EPI_ISL_649127|2020-0 ···················· ···················· ···················· ···················· ···················· ····················
Q 5360 hCoV-19/Japan/TKYE622387/2020|EPI_ISL_649128|2020- ···················· ···················· ···················· ···················· ···················· ····················
Q 5361 hCoV-19/Japan/TKYE622388/2020|EPI_ISL_649129|2020- ···················· ···················· ···················· ···················· ···················· ····················
Q 5362 hCoV-19/Japan/TKYE622391/2020|EPI_ISL_649130|2020- ···················· ···················· ···················· ···················· ···················· ····················
Q 5363 hCoV-19/Japan/TKYE622393/2020|EPI_ISL_649131|2020- ···················· ···················· ···················· ···················· ···················· ····················
Q 5364 hCoV-19/Japan/TKYT22395/2020|EPI_ISL_649132|2020-0 ···················· ···················· ···················· ···················· ···················· ····················
Q 5365 hCoV-19/Japan/TKYE622411/2020|EPI_ISL_649133|2020- ···················· ···················· ···················· ···················· ···················· ····················
Q 5366 hCoV-19/Japan/TKYE622414/2020|EPI_ISL_649134|2020- ···················· ···················· ···················· ···················· ···················· ····················
Q 5367 hCoV-19/Japan/TKYE624705/2020|EPI_ISL_649135|2020- ···················· ···················· ···················· ···················· ···················· ····················
Q 5368 hCoV-19/Japan/TKYE627000/2020|EPI_ISL_649136|2020- ···················· ···················· ···················· ···················· ···················· ····················
Q 5369 hCoV-19/Japan/TKYET29933/2020|EPI_ISL_649137|2020- ···················· ···················· ···················· ···················· ···················· ····················
Q 5370 hCoV-19/Japan/TKYE630074/2020|EPI_ISL_649138|2020- ···················· ···················· ···················· ···················· ···················· ····················
Q 5371 hCoV-19/Japan/TKYE630094/2020|EPI_ISL_649139|2020- ···················· ···················· ···················· ···················· ···················· ····················
Q 5372 hCoV-19/Japan/TKYT30095/2020|EPI_ISL_649140|2020-0 ···················· ···················· ···················· ···················· ···················· ····················
Q 5373 hCoV-19/Japan/TKYT30408/2020|EPI_ISL_649141|2020-0 ···················· ···················· ···················· ···················· ···················· ····················
Q 5374 hCoV-19/Japan/TKYE630411/2020|EPI_ISL_649142|2020- ···················· ···················· ···················· ···················· ···················· ····················
Q 5375 hCoV-19/Japan/TKYE630414/2020|EPI_ISL_649143|2020- ···················· ···················· ···················· ···················· ···················· ····················
Q 5376 hCoV-19/Japan/TKYT30430/2020|EPI_ISL_649144|2020-0 ···················· ···················· ···················· ···················· ···················· ····················
Q 5377 hCoV-19/England/NOTT-118FDA/2020|EPI_ISL_627700|20 ···················· ···················· ···················· ···················· ···················· ····················
Q 5378 hCoV-19/England/NOTT-118FF8/2020|EPI_ISL_627701|20 ···················· ···················· ···················· ···················· ···················· ····················
Q 5379 hCoV-19/England/NOTT-119003/2020|EPI_ISL_627702|20 ···················· ···················· ···················· ···················· ···················· ····················
Q 5380 hCoV-19/England/NOTT-11906D/2020|EPI_ISL_627703|20 ···················· ···················· ···················· ···················· ···················· ····················
Q 5381 hCoV-19/England/NOTT-11908B/2020|EPI_ISL_627704|20 ···················· ···················· ···················· ···················· ···················· ····················
Q 5382 hCoV-19/England/NOTT-11909A/2020|EPI_ISL_627705|20 ···················· ···················· ···················· ···················· ···················· ····················
Q 5383 hCoV-19/USA/NY-NYCPHL-000823/2020|EPI_ISL_632200|2 ···················· ···················· ···················· ···················· ···················· ····················
Q 5384 hCoV-19/USA/NY-NYCPHL-000824/2020|EPI_ISL_632201|2 ···················· ···················· ···················· ···················· ···················· ····················
Q 5385 hCoV-19/USA/NY-NYCPHL-000825/2020|EPI_ISL_632202|2 ···················· ···················· ···················· ···················· ···················· ····················
Q 5386 hCoV-19/USA/NY-NYCPHL-000826/2020|EPI_ISL_632203|2 ···················· ···················· ···················· ···················· ···················· ····················
Q 5387 hCoV-19/USA/NY-NYCPHL-000827/2020|EPI_ISL_632204|2 ···················· ···················· ···················· ···················· ···················· ····················
Q 5388 hCoV-19/USA/NY-NYCPHL-000828/2020|EPI_ISL_632205|2 ···················· ···················· ···················· ···················· ···················· ····················
Q 5389 hCoV-19/USA/NY-NYCPHL-000829/2020|EPI_ISL_632206|2 ···················· ···················· ···················· ···················· ···················· ····················
Q 5390 hCoV-19/USA/NY-NYCPHL-000830/2020|EPI_ISL_632207|2 ···················· ···················· ···················· ···················· ···················· ····················
Q 5391 hCoV-19/USA/NY-NYCPHL-000831/2020|EPI_ISL_632208|2 ···················· ···················· ···················· ···················· ···················· ····················
Q 5392 hCoV-19/USA/NY-NYCPHL-000832/2020|EPI_ISL_632209|2 ···················· ···················· ···················· ···················· ···················· ····················
Q 5393 hCoV-19/USA/NY-NYCPHL-000833/2020|EPI_ISL_632210|2 ···················· ···················· ···················· ···················· ···················· ····················
Q 5394 hCoV-19/USA/NY-NYCPHL-000834/2020|EPI_ISL_632211|2 ···················· ···················· ···················· ···················· ···················· ····················
Q 5395 hCoV-19/USA/NY-NYCPHL-000835/2020|EPI_ISL_632212|2 ···················· ···················· ···················· ···················· ···················· ····················
Q 5396 hCoV-19/USA/NY-NYCPHL-000836/2020|EPI_ISL_632213|2 ···················· ···················· ···················· ···················· ···················· ····················
Q 5397 hCoV-19/USA/NY-NYCPHL-000837/2020|EPI_ISL_632214|2 ···················· ···················· ···················· ···················· ···················· ····················
Q 5398 hCoV-19/USA/NY-NYCPHL-000838/2020|EPI_ISL_632215|2 ···················· ···················· ···················· ···················· ···················· ····················
Q 5399 hCoV-19/USA/NY-NYCPHL-000839/2020|EPI_ISL_632216|2 ···················· ···················· ···················· ···················· ···················· ····················
Q 5400 hCoV-19/USA/NY-NYCPHL-000840/2020|EPI_ISL_632217|2 ···················· ···················· ···················· ···················· ···················· ····················
Q 5401 hCoV-19/USA/NY-NYCPHL-000841/2020|EPI_ISL_632218|2 ···················· ···················· ···················· ···················· ···················· ····················
Q 5402 hCoV-19/USA/NY-NYCPHL-000842/2020|EPI_ISL_632219|2 ···················· ···················· ···················· ···················· ···················· ····················
Q 5403 hCoV-19/USA/NY-NYCPHL-000843/2020|EPI_ISL_632220|2 ···················· ···················· ···················· ···················· ···················· ····················
Q 5404 hCoV-19/USA/NY-NYCPHL-000844/2020|EPI_ISL_632221|2 ···················· ···················· ···················· ···················· ···················· ····················
Q 5405 hCoV-19/USA/NY-NYCPHL-000845/2020|EPI_ISL_632222|2 ···················· ···················· ···················· ···················· ···················· ····················
Q 5406 hCoV-19/USA/NY-NYCPHL-000846/2020|EPI_ISL_632223|2 ···················· ···················· ···················· ···················· ···················· ····················
Q 5407 hCoV-19/USA/NY-NYCPHL-000847/2020|EPI_ISL_632224|2 ···················· ···················· ···················· ···················· ···················· ····················
Q 5408 hCoV-19/USA/NY-NYCPHL-000848/2020|EPI_ISL_632225|2 ···················· ···················· ···················· ···················· ···················· ····················
Q 5409 hCoV-19/USA/NY-NYCPHL-000849/2020|EPI_ISL_632226|2 ···················· ···················· ···················· ···················· ···················· ····················
Q 5410 hCoV-19/USA/NY-NYCPHL-000850/2020|EPI_ISL_632227|2 ···················· ···················· ···················· ···················· ···················· ····················
Q 5411 hCoV-19/USA/NY-NYCPHL-000851/2020|EPI_ISL_632228|2 ···················· ···················· ···················· ···················· ···················· ····················
Q 5412 hCoV-19/USA/NY-NYCPHL-000852/2020|EPI_ISL_632229|2 ···················· ···················· ···················· ···················· ···················· ····················
Q 5413 hCoV-19/USA/NY-NYCPHL-000854/2020|EPI_ISL_632230|2 ···················· ···················· ···················· ···················· ···················· ····················
Q 5414 hCoV-19/USA/NY-NYCPHL-000855/2020|EPI_ISL_632231|2 ···················· ···················· ···················· ···················· ···················· ····················
Q 5415 hCoV-19/USA/NY-NYCPHL-000856/2020|EPI_ISL_632232|2 ···················· ···················· ···················· ···················· ···················· ····················
Q 5416 hCoV-19/USA/NY-NYCPHL-000857/2020|EPI_ISL_632233|2 ···················· ···················· ···················· ···················· ···················· ····················
Q 5417 hCoV-19/USA/NY-NYCPHL-000858/2020|EPI_ISL_632234|2 ···················· ···················· ···················· ···················· ···················· ····················
Q 5418 hCoV-19/USA/NY-NYCPHL-000859/2020|EPI_ISL_632235|2 ···················· ···················· ···················· ···················· ···················· ····················
Q 5419 hCoV-19/USA/NY-NYCPHL-000861/2020|EPI_ISL_632237|2 ···················· ···················· ···················· ···················· ···················· ····················
Q 5420 hCoV-19/USA/NY-NYCPHL-000864/2020|EPI_ISL_632240|2 ···················· ···················· ···················· ···················· ···················· ····················
Q 5421 hCoV-19/Scotland/GCVR-16FC91/2020|EPI_ISL_632247|2 ···················· ···················· ···················· ···················· ···················· ····················
Q 5422 hCoV-19/Scotland/GCVR-16FD70/2020|EPI_ISL_632248|2 ···················· ·A·················· ···················· ···················· ···················· ····················
Q 5423 hCoV-19/Scotland/GCVR-16F97C/2020|EPI_ISL_632249|2 ···················· ·A·················· ···················· ···················· ···················· ····················
Q 5424 hCoV-19/Bahrain/340860543/2020|EPI_ISL_632250|2020 ···················· ···················· ···················· ···················· ···················· ····················
Q 5425 hCoV-19/Bahrain/340860476/2020|EPI_ISL_632251|2020 ···················· ···················· ···················· ···················· ···················· ····················
Q 5426 hCoV-19/Bahrain/340861142/2020|EPI_ISL_632252|2020 ···················· ···················· ···················· ···················· ···················· ····················

| | | | | | | | | |
|---|---|---|---|---|---|---|---|---|
| Q | 5427 | hCoV-19/Bahrain/340861254_S20/2020\|EPI_ISL_632253\| | ···················· | ···················· | ···················· | ···················· | ···················· | ···················· |
| Q | 5428 | hCoV-19/Bahrain/340861245_S21/2020\|EPI_ISL_632254\| | ···················· | ···················· | ···················· | ···················· | ···················· | ···················· |
| Q | 5429 | hCoV-19/Bahrain/340861247_S22/2020\|EPI_ISL_632255\| | ···················· | ···················· | ···················· | ···················· | ···················· | ···················· |
| Q | 5430 | hCoV-19/Bahrain/340861274_S26/2020\|EPI_ISL_632256\| | ···················· | ···················· | ···················· | ···················· | ···················· | ···················· |
| Q | 5431 | hCoV-19/Bahrain/340861272_S28/2020\|EPI_ISL_632257\| | ···················· | ···················· | ···················· | ···················· | ···················· | ···················· |
| Q | 5432 | hCoV-19/Bahrain/600023154_S29/2020\|EPI_ISL_632258\| | ···················· | ···················· | ···················· | ···················· | ···················· | ···················· |
| Q | 5433 | hCoV-19/Bahrain/340864186/2020\|EPI_ISL_632259\|2020 | ···················· | ···················· | ···················· | ···················· | ···················· | ···················· |
| Q | 5434 | hCoV-19/Bahrain/340861258/2020\|EPI_ISL_632260\|2020 | ···················· | ···················· | ···················· | ···················· | ···················· | ···················· |
| Q | 5435 | hCoV-19/Bahrain/340286642_S20_L001/2020\|EPI_ISL_63 | ···················· | ···················· | ···················· | ···················· | ···················· | ···················· |
| Q | 5436 | hCoV-19/Bahrain/340513774_S1_L001/2020\|EPI_ISL_632 | ···················· | ···················· | ···················· | ···················· | ···················· | ···················· |
| Q | 5437 | hCoV-19/Bahrain/340564387_S8_L001/2020\|EPI_ISL_632 | ···················· | ···················· | ···················· | ···················· | ···················· | ···················· |
| Q | 5438 | hCoV-19/Bahrain/340809497_S11_L001/2020\|EPI_ISL_63 | ···················· | ···················· | ···················· | ···················· | ···················· | ···················· |
| Q | 5439 | hCoV-19/Bahrain/920262933_S19_L001/2020\|EPI_ISL_63 | ···················· | ···················· | ···················· | ···················· | ···················· | ···················· |
| Q | 5440 | hCoV-19/Bahrain/920265546_S18_L001/2020\|EPI_ISL_63 | ···················· | ···················· | ···················· | ···················· | ···················· | ··A················· |
| Q | 5441 | hCoV-19/Bahrain/920319237_S17_L001/2020\|EPI_ISL_63 | ···················· | ···················· | ···················· | ···················· | ···················· | ··A················· |
| Q | 5442 | hCoV-19/Bahrain/340864151_S32/2020\|EPI_ISL_632268\| | ···················· | ···················· | ···················· | ···················· | ···················· | ···················· |
| Q | 5443 | hCoV-19/Bahrain/340858468_S5/2020\|EPI_ISL_632269\|2 | ···················· | ···················· | ···················· | ···················· | ···················· | ···················· |
| Q | 5444 | hCoV-19/Bahrain/340859154_S6/2020\|EPI_ISL_632270\|2 | ···················· | ···················· | ···················· | ···················· | ···················· | ···················· |
| Q | 5445 | hCoV-19/Bahrain/340859160_S7/2020\|EPI_ISL_632271\|2 | ···················· | ···················· | ···················· | ···················· | ···················· | ···················· |
| Q | 5446 | hCoV-19/Bahrain/340859209_S2/2020\|EPI_ISL_632272\|2 | ···················· | ···················· | ···················· | ···················· | ···················· | ···················· |
| Q | 5447 | hCoV-19/Bahrain/340859251_S1/2020\|EPI_ISL_632273\|2 | ···················· | ···················· | ···················· | ···················· | ···················· | ···················· |
| Q | 5448 | hCoV-19/Bahrain/340859839_S13/2020\|EPI_ISL_632274\| | ···················· | ···················· | ···················· | ···················· | ···················· | ···················· |
| Q | 5449 | hCoV-19/Bahrain/340859842_S14/2020\|EPI_ISL_632275\| | ···················· | ···················· | ···················· | ···················· | ···················· | ···················· |
| Q | 5450 | hCoV-19/Bahrain/340859867_S9/2020\|EPI_ISL_632276\|2 | ···················· | ···················· | ···················· | ···················· | ···················· | ···················· |
| Q | 5451 | hCoV-19/Bahrain/340859906_S10/2020\|EPI_ISL_632277\| | ···················· | ···················· | ···················· | ···················· | ···················· | ···················· |
| Q | 5452 | hCoV-19/Bahrain/340859913_S11/2020\|EPI_ISL_632278\| | ···················· | ···················· | ···················· | ···················· | ···················· | ···················· |
| Q | 5453 | hCoV-19/Bahrain/340859926_S12/2020\|EPI_ISL_632279\| | ···················· | ···················· | ···················· | ···················· | ···················· | ···················· |
| Q | 5454 | hCoV-19/Bahrain/340860476_S15/2020\|EPI_ISL_632280\| | ···················· | ···················· | ···················· | ···················· | ···················· | ···················· |
| Q | 5455 | hCoV-19/Bahrain/340860489_S16/2020\|EPI_ISL_632281\| | ···················· | ···················· | ···················· | ···················· | ···················· | ···················· |
| Q | 5456 | hCoV-19/Bahrain/340860526_S17/2020\|EPI_ISL_632282\| | ···················· | ···················· | ···················· | ···················· | ···················· | ···················· |
| Q | 5457 | hCoV-19/USA/FL-UF-29/2020\|EPI_ISL_632287\|2020-07-2 | ···················· | ···················· | ···················· | ···················· | ···················· | ···················· |
| Q | 5458 | hCoV-19/USA/MA-JLL-17S/2020\|EPI_ISL_632288\|2020 | ···················· | ···················· | ···················· | ···················· | ···················· | ···················· |
| Q | 5459 | hCoV-19/USA/MA-JLL-4O/2020\|EPI_ISL_632289\|2020 | ···················· | ···················· | ···················· | ···················· | ···················· | ···················· |
| Q | 5460 | hCoV-19/USA/MA-JLL-26N/2020\|EPI_ISL_632290\|2020 | ···················· | ···················· | ···················· | ···················· | ···················· | ···················· |
| Q | 5461 | hCoV-19/USA/MA-JLL-26O/2020\|EPI_ISL_632291\|2020 | ···················· | ···················· | ···················· | ···················· | ···················· | ···················· |
| Q | 5462 | hCoV-19/USA/MA-JLL-26S/2020\|EPI_ISL_632292\|2020 | ···················· | ···················· | ···················· | ···················· | ···················· | ···················· |
| Q | 5463 | hCoV-19/USA/MA-JLL-145N/2020\|EPI_ISL_632293\|2020 | ···················· | ···················· | ···················· | ···················· | ···················· | ···················· |
| Q | 5464 | hCoV-19/USA/MA-JLL-147N/2020\|EPI_ISL_632294\|2020 | ···················· | ···················· | ···················· | ···················· | ···················· | ···················· |
| Q | 5465 | hCoV-19/USA/MA-JLL-149N/2020\|EPI_ISL_632295\|2020 | ···················· | ···················· | ···················· | ···················· | ···················· | ···················· |
| Q | 5466 | hCoV-19/USA/MA-JLL-150N/2020\|EPI_ISL_632296\|2020 | ···················· | ···················· | ···················· | ···················· | ···················· | ···················· |
| Q | 5467 | hCoV-19/USA/MA-JLL-154/2020\|EPI_ISL_632297\|2020 | ···················· | ···················· | ···················· | ···················· | ···················· | ···················· |
| Q | 5468 | hCoV-19/USA/MA-JLL-155N/2020\|EPI_ISL_632298\|2020 | ···················· | ···················· | ···················· | ···················· | ···················· | ···················· |
| Q | 5469 | hCoV-19/USA/MA-JLL-159N/2020\|EPI_ISL_632299\|2020 | ···················· | ···················· | ···················· | ···················· | ···················· | ···················· |
| Q | 5470 | hCoV-19/England/QEUH-AE2FD0/2020\|EPI_ISL_633500\|20 | ···················· | ···················· | ···················· | ···················· | ···················· | ···················· |
| Q | 5471 | hCoV-19/England/QEUH-AE287B/2020\|EPI_ISL_633501\|20 | ···················· | ···················· | ···················· | ···················· | ···················· | ···················· |
| Q | 5472 | hCoV-19/England/QEUH-AE1CAD/2020\|EPI_ISL_633502\|20 | ···················· | ···················· | ···················· | ···················· | ···················· | ···················· |
| Q | 5473 | hCoV-19/England/QEUH-AE218F/2020\|EPI_ISL_633503\|20 | ···················· | ···················· | ···················· | ···················· | ···················· | ···················· |
| Q | 5474 | hCoV-19/England/QEUH-AE2468/2020\|EPI_ISL_633504\|20 | ···················· | ···················· | ···················· | ···················· | ···················· | ···················· |
| Q | 5475 | hCoV-19/England/QEUH-AE2055/2020\|EPI_ISL_633505\|20 | ···················· | ···················· | ···················· | ···················· | ···················· | ···················· |
| Q | 5476 | hCoV-19/England/QEUH-AE284E/2020\|EPI_ISL_633506\|20 | ···················· | ···················· | ···················· | ···················· | ···················· | ···················· |
| Q | 5477 | hCoV-19/England/QEUH-AE1C25/2020\|EPI_ISL_633507\|20 | ···················· | ···················· | ···················· | ···················· | ···················· | ···················· |
| Q | 5478 | hCoV-19/England/QEUH-AE20BF/2020\|EPI_ISL_633508\|20 | ···················· | ···················· | ···················· | ···················· | ···················· | ···················· |
| Q | 5479 | hCoV-19/England/QEUH-AE29F0/2020\|EPI_ISL_633509\|20 | ···················· | ···················· | ···················· | ···················· | ···················· | ···················· |
| Q | 5480 | hCoV-19/England/QEUH-AE3160/2020\|EPI_ISL_633510\|20 | ···················· | ···················· | ···················· | ···················· | ···················· | ···················· |
| Q | 5481 | hCoV-19/England/QEUH-AE1D40/2020\|EPI_ISL_633511\|20 | ···················· | ···················· | ···················· | ···················· | ···················· | ···················· |
| Q | 5482 | hCoV-19/England/QEUH-AE25A1/2020\|EPI_ISL_633512\|20 | ···················· | ···················· | ···················· | ···················· | ···················· | ···················· |
| Q | 5483 | hCoV-19/England/QEUH-AE21E9/2020\|EPI_ISL_633513\|20 | ···················· | ···················· | ···················· | ···················· | ···················· | ···················· |
| Q | 5484 | hCoV-19/England/QEUH-AE2EE2/2020\|EPI_ISL_633515\|20 | ···················· | ···················· | ···················· | ···················· | ···················· | ···················· |
| Q | 5485 | hCoV-19/England/QEUH-AE1BCE/2020\|EPI_ISL_633525\|20 | ···················· | ···················· | ···················· | ···················· | ···················· | ···················· |
| Q | 5486 | hCoV-19/England/QEUH-AE1C52/2020\|EPI_ISL_633526\|20 | ···················· | ···················· | ···················· | ···················· | ···················· | ···················· |
| Q | 5487 | hCoV-19/England/QEUH-AE1EC5/2020\|EPI_ISL_633527\|20 | ···················· | ···················· | ···················· | ···················· | ···················· | ···················· |
| Q | 5488 | hCoV-19/England/QEUH-AE2A66/2020\|EPI_ISL_633528\|20 | ···················· | ···················· | ···················· | ···················· | ···················· | ···················· |
| Q | 5489 | hCoV-19/England/QEUH-AE1ED4/2020\|EPI_ISL_633529\|20 | ···················· | ···················· | ···················· | ···················· | ···················· | ···················· |
| Q | 5490 | hCoV-19/England/QEUH-AE2C8E/2020\|EPI_ISL_633530\|20 | ···················· | ···················· | ···················· | ···················· | ···················· | ···················· |
| Q | 5491 | hCoV-19/England/QEUH-AE1B19/2020\|EPI_ISL_633531\|20 | ···················· | ···················· | ···················· | ···················· | ···················· | ···················· |
| Q | 5492 | hCoV-19/England/QEUH-AE3027/2020\|EPI_ISL_633532\|20 | ···················· | ···················· | ···················· | ···················· | ···················· | ···················· |
| Q | 5493 | hCoV-19/England/QEUH-AE1F1D/2020\|EPI_ISL_633533\|20 | ···················· | ···················· | ···················· | ···················· | ···················· | ···················· |
| Q | 5494 | hCoV-19/England/QEUH-AE26DB/2020\|EPI_ISL_633534\|20 | ···················· | ···················· | ···················· | ···················· | ···················· | ···················· |
| Q | 5495 | hCoV-19/England/QEUH-AE3115/2020\|EPI_ISL_633535\|20 | ···················· | ···················· | ···················· | ···················· | ···················· | ···················· |
| Q | 5496 | hCoV-19/Wales/QEUH-AE23D4/2020\|EPI_ISL_633536\|2020 | ···················· | ···················· | ···················· | ···················· | ···T················ | ···················· |
| Q | 5497 | hCoV-19/England/QEUH-AE29A5/2020\|EPI_ISL_633537\|20 | ···················· | ···················· | ···················· | ···················· | ···················· | ···················· |
| Q | 5498 | hCoV-19/England/QEUH-AE2547/2020\|EPI_ISL_633538\|20 | ···················· | ···················· | ···················· | ···················· | ···················· | ···················· |
| Q | 5499 | hCoV-19/England/QEUH-AE24FF/2020\|EPI_ISL_633539\|20 | ···················· | ···················· | ···················· | ···················· | ···················· | ···················· |
| Q | 5500 | hCoV-19/England/QEUH-AE24D1/2020\|EPI_ISL_633541\|20 | ···················· | ···················· | ···················· | ···················· | ···················· | ···················· |
| Q | 5501 | hCoV-19/England/QEUH-AE2FC1/2020\|EPI_ISL_633542\|20 | ···················· | ···················· | ···················· | ···················· | ···················· | ···················· |
| Q | 5502 | hCoV-19/England/QEUH-AE1FE0/2020\|EPI_ISL_633543\|20 | ···················· | ···················· | ···················· | ···················· | ···················· | ···················· |
| Q | 5503 | hCoV-19/England/QEUH-AE3045/2020\|EPI_ISL_633544\|20 | ···················· | ···················· | ···················· | ···················· | ···················· | ···················· |
| Q | 5504 | hCoV-19/England/QEUH-AE23E3/2020\|EPI_ISL_633545\|20 | ···················· | ···················· | ···················· | ···················· | ···················· | ···················· |
| Q | 5505 | hCoV-19/England/QEUH-AE1A85/2020\|EPI_ISL_633546\|20 | ···················· | ···················· | ···················· | ···················· | ···················· | ···················· |
| Q | 5506 | hCoV-19/England/QEUH-AE1AA3/2020\|EPI_ISL_633547\|20 | ···················· | ···················· | ···················· | ···················· | ···················· | ···················· |

| | | | | | | | |
|---|---|---|---|---|---|---|---|
| Q | 5507 hCoV-19/England/QEUH-AE2AB1/2020\|EPI_ISL_633548\|20 | ···················· | ·A·················· | ····················· | ···················· | ···················· | ···················· |
| Q | 5508 hCoV-19/England/QEUH-AE294B/2020\|EPI_ISL_633549\|20 | ···················· | ···················· | ····················· | ···················· | ···················· | ···················· |
| Q | 5509 hCoV-19/England/QEUH-AE286C/2020\|EPI_ISL_633550\|20 | ···················· | ···················· | ····················· | ···················· | ···················· | ···················· |
| Q | 5510 hCoV-19/England/QEUH-AE2B81/2020\|EPI_ISL_633551\|20 | ···················· | ···················· | ····················· | ···················· | ···················· | ···················· |
| Q | 5511 hCoV-19/England/QEUH-AE251A/2020\|EPI_ISL_633552\|20 | ···················· | ···················· | ····················· | ···················· | ···················· | ···················· |
| Q | 5512 hCoV-19/England/QEUH-AE1AFE/2020\|EPI_ISL_633553\|20 | ···················· | ···················· | ····················· | ···················· | ···················· | ···················· |
| Q | 5513 hCoV-19/England/QEUH-AE2389/2020\|EPI_ISL_633554\|20 | ···················· | ···················· | ····················· | ···················· | ···················· | ···················· |
| Q | 5514 hCoV-19/England/QEUH-AE1BFB/2020\|EPI_ISL_633555\|20 | ···················· | ···················· | ····················· | ···················· | ···················· | ···················· |
| Q | 5515 hCoV-19/England/QEUH-AE3018/2020\|EPI_ISL_633556\|20 | ···················· | ···················· | ····················· | ···················· | ···················· | ···················· |
| Q | 5516 hCoV-19/England/QEUH-AE1CDA/2020\|EPI_ISL_633557\|20 | ···················· | ···················· | ····················· | ···················· | ···················· | ···················· |
| Q | 5517 hCoV-19/England/QEUH-AE2C60/2020\|EPI_ISL_633558\|20 | ···················· | ···················· | ····················· | ···················· | ···················· | ···················· |
| Q | 5518 hCoV-19/England/QEUH-AE2CBB/2020\|EPI_ISL_633559\|20 | ···················· | ···················· | ····················· | ···················· | ···················· | ···················· |
| Q | 5519 hCoV-19/England/QEUH-AE1FD1/2020\|EPI_ISL_633560\|20 | ···················· | ···················· | ····················· | ···················· | ···················· | ···················· |
| Q | 5520 hCoV-19/England/QEUH-AE2D21/2020\|EPI_ISL_633561\|20 | ···················· | ···················· | ····················· | ···················· | ···················· | ···················· |
| Q | 5521 hCoV-19/England/QEUH-AE1A49/2020\|EPI_ISL_633562\|20 | ···················· | ···················· | ····················· | ···················· | ···················· | ···················· |
| Q | 5522 hCoV-19/England/QEUH-AE225F/2020\|EPI_ISL_633563\|20 | ···················· | ···················· | ····················· | ···················· | ···················· | ···················· |
| Q | 5523 hCoV-19/England/QEUH-AE277E/2020\|EPI_ISL_633564\|20 | ···················· | ···················· | ····················· | ···················· | ···················· | ···················· |
| Q | 5524 hCoV-19/England/QEUH-AE2170/2020\|EPI_ISL_633565\|20 | ···················· | ···················· | ····················· | ···················· | ···················· | ···················· |
| Q | 5525 hCoV-19/England/QEUH-AE2F0D/2020\|EPI_ISL_633566\|20 | ···················· | ···················· | ····················· | ···················· | ···················· | ···················· |
| Q | 5526 hCoV-19/England/QEUH-AE2FA3/2020\|EPI_ISL_633570\|20 | ···················· | ···················· | ····················· | ···················· | ···················· | ···················· |
| Q | 5527 hCoV-19/England/QEUH-AE2899/2020\|EPI_ISL_633571\|20 | ···················· | ···················· | ····················· | ···················· | ···················· | ···················· |
| Q | 5528 hCoV-19/England/QEUH-AE279C/2020\|EPI_ISL_633578\|20 | ···················· | ···················· | ····················· | ···················· | ···················· | ···················· |
| Q | 5529 hCoV-19/England/QEUH-AE25DE/2020\|EPI_ISL_633579\|20 | ···················· | ···················· | ····················· | ···················· | ···················· | ···················· |
| Q | 5530 hCoV-19/England/QEUH-AE3133/2020\|EPI_ISL_633580\|20 | ···················· | ···················· | ····················· | ···················· | ···················· | ···················· |
| Q | 5531 hCoV-19/England/QEUH-AE24C2/2020\|EPI_ISL_633581\|20 | ···················· | ···················· | ····················· | ···················· | ···················· | ···················· |
| Q | 5532 hCoV-19/England/QEUH-AE2486/2020\|EPI_ISL_633582\|20 | ···················· | ···················· | ····················· | ···················· | ···················· | ···················· |
| Q | 5533 hCoV-19/England/QEUH-AE2A93/2020\|EPI_ISL_633583\|20 | ···················· | ···················· | ····················· | ···················· | ···················· | ···················· |
| Q | 5534 hCoV-19/England/QEUH-AE2C42/2020\|EPI_ISL_633584\|20 | ···················· | ···················· | ····················· | ···················· | ···················· | ···················· |
| Q | 5535 hCoV-19/England/QEUH-AE1CBC/2020\|EPI_ISL_633586\|20 | ···················· | ···················· | ····················· | ···················· | ···················· | ···················· |
| Q | 5536 hCoV-19/England/QEUH-AE2C7F/2020\|EPI_ISL_633587\|20 | ···················· | ···················· | ····················· | ···················· | ···················· | ···················· |
| Q | 5537 hCoV-19/England/QEUH-AE1DC8/2020\|EPI_ISL_633588\|20 | ···················· | ···················· | ····················· | ···················· | ···················· | ···················· |
| Q | 5538 hCoV-19/England/QEUH-AE2F76/2020\|EPI_ISL_633591\|20 | ···················· | ···················· | ····················· | ···················· | ···················· | ···················· |
| Q | 5539 hCoV-19/England/QEUH-AE2064/2020\|EPI_ISL_633592\|20 | ···················· | ···················· | ····················· | ···················· | ···················· | ···················· |
| Q | 5540 hCoV-19/England/QEUH-AE21AD/2020\|EPI_ISL_633593\|20 | ···················· | ···················· | ····················· | ···················· | ···················· | ···················· |
| Q | 5541 hCoV-19/England/QEUH-AE2B90/2020\|EPI_ISL_633594\|20 | ···················· | ···················· | ····················· | ···················· | ···················· | ···················· |
| Q | 5542 hCoV-19/England/QEUH-AE1B82/2020\|EPI_ISL_633595\|20 | ···················· | ···················· | ····················· | ···················· | ···················· | ···················· |
| Q | 5543 hCoV-19/England/QEUH-AE2213/2020\|EPI_ISL_633596\|20 | ···················· | ···················· | ····················· | ···················· | ···················· | ···················· |
| Q | 5544 hCoV-19/England/QEUH-AE2459/2020\|EPI_ISL_633597\|20 | ···················· | ···················· | ····················· | ···················· | ···················· | ···················· |
| Q | 5545 hCoV-19/England/QEUH-AE23B6/2020\|EPI_ISL_633598\|20 | ···················· | ···················· | ····················· | ···················· | ···················· | ···················· |
| Q | 5546 hCoV-19/England/QEUH-AE1BBF/2020\|EPI_ISL_633599\|20 | ···················· | ···················· | ····················· | ···················· | ···················· | ···················· |
| Q | 5547 hCoV-19/Wales/CAMC-AAF889/2020\|EPI_ISL_634800\|2020 | ···················· | ···················· | ····················· | ···················· | ···················· | ···················· |
| Q | 5548 hCoV-19/Wales/CAMC-AAFD99/2020\|EPI_ISL_634801\|2020 | ···················· | ···················· | ····················· | ···················· | ···················· | ···················· |
| Q | 5549 hCoV-19/Wales/CAMC-AAFBBD/2020\|EPI_ISL_634802\|2020 | ···················· | ···················· | ····················· | ···················· | ···················· | ···················· |
| Q | 5550 hCoV-19/Wales/CAMC-AAFBAE/2020\|EPI_ISL_634803\|2020 | ···················· | ···················· | ····················· | ···················· | ···················· | ···················· |
| Q | 5551 hCoV-19/Wales/CAMC-AAFBDB/2020\|EPI_ISL_634804\|2020 | ···················· | ···················· | ····················· | ···················· | ···················· | ···················· |
| Q | 5552 hCoV-19/Wales/CAMC-AAFB9F/2020\|EPI_ISL_634805\|2020 | ···················· | ···················· | ····················· | ···················· | ···················· | ···················· |
| Q | 5553 hCoV-19/Wales/CAMC-AAFBCC/2020\|EPI_ISL_634806\|2020 | ···················· | ···················· | ····················· | ···················· | ···················· | ···················· |
| Q | 5554 hCoV-19/Wales/CAMC-AAFDA8/2020\|EPI_ISL_634807\|2020 | ···················· | ···················· | ····················· | ···················· | ···················· | ···················· |
| Q | 5555 hCoV-19/Wales/CAMC-AB1BF4/2020\|EPI_ISL_634808\|2020 | ···················· | ···················· | ····················· | ···················· | ···················· | ···················· |
| Q | 5556 hCoV-19/Wales/CAMC-AB1FF8/2020\|EPI_ISL_634809\|2020 | ···················· | ···················· | ····················· | ···················· | ···················· | ···················· |
| Q | 5557 hCoV-19/Wales/CAMC-AB23A0/2020\|EPI_ISL_634810\|2020 | ···················· | ···················· | ····················· | ···················· | ···················· | ···················· |
| Q | 5558 hCoV-19/Wales/CAMC-AB23BF/2020\|EPI_ISL_634811\|2020 | ···················· | ···················· | ····················· | ···················· | ···················· | ···················· |
| Q | 5559 hCoV-19/Wales/CAMC-AB2030/2020\|EPI_ISL_634812\|2020 | ···················· | ···················· | ····················· | ···················· | ···················· | ···················· |
| Q | 5560 hCoV-19/Wales/CAMC-AB213D/2020\|EPI_ISL_634813\|2020 | ···················· | ···················· | ····················· | ···················· | ···················· | ···················· |
| Q | 5561 hCoV-19/Wales/CAMC-AB182A/2020\|EPI_ISL_634814\|2020 | ···················· | ···················· | ····················· | ···················· | ···················· | ···················· |
| Q | 5562 hCoV-19/Wales/CAMC-AB2391/2020\|EPI_ISL_634815\|2020 | ···················· | ···················· | ····················· | ···················· | ···················· | ···················· |
| Q | 5563 hCoV-19/Wales/CAMC-AB212E/2020\|EPI_ISL_634816\|2020 | ···················· | ···················· | ····················· | ···················· | ···················· | ···················· |
| Q | 5564 hCoV-19/France/ARA-SC147/2020\|EPI_ISL_634817\|2020- | ···················· | ···················· | ····················· | ···················· | ···················· | ···················· |
| Q | 5565 hCoV-19/France/ARA-SC150/2020\|EPI_ISL_634818\|2020- | ···················· | ···················· | ····················· | ···················· | ···················· | ···················· |
| Q | 5566 hCoV-19/France/ARA-SC151/2020\|EPI_ISL_634819\|2020- | ···················· | ···················· | ····················· | ···················· | ···················· | ···················· |
| Q | 5567 hCoV-19/France/ARA-SC152/2020\|EPI_ISL_634820\|2020- | ···················· | ···················· | ····················· | ···················· | ···················· | ···················· |
| Q | 5568 hCoV-19/France/ARA-SC153/2020\|EPI_ISL_634821\|2020- | ···················· | ···················· | ····················· | ···················· | ···················· | ···················· |
| Q | 5569 hCoV-19/France/ARA-SC154/2020\|EPI_ISL_634822\|2020- | ···················· | ···················· | ····················· | ···················· | ···················· | ···················· |
| Q | 5570 hCoV-19/France/ARA-SC156/2020\|EPI_ISL_634823\|2020- | ···················· | ···················· | ····················· | ···················· | ···················· | ···················· |
| Q | 5571 hCoV-19/France/ARA-SC159/2020\|EPI_ISL_634824\|2020- | ···················· | ···················· | ····················· | ···················· | ···················· | ···················· |
| Q | 5572 hCoV-19/France/ARA-SC160/2020\|EPI_ISL_634825\|2020- | ···················· | ···················· | ····················· | ···················· | ···················· | ···················· |
| Q | 5573 hCoV-19/France/ARA-SC161/2020\|EPI_ISL_634826\|2020- | ···················· | ···················· | ····················· | ···················· | ···················· | ···················· |
| Q | 5574 hCoV-19/France/ARA-SC162/2020\|EPI_ISL_634827\|2020- | ···················· | ···················· | ····················· | ···················· | ···················· | ···················· |
| Q | 5575 hCoV-19/France/ARA-SC167/2020\|EPI_ISL_634828\|2020- | ···················· | ···················· | ····················· | ···················· | ···················· | ···················· |
| Q | 5576 hCoV-19/France/ARA-SC168/2020\|EPI_ISL_634829\|2020- | ···················· | ···················· | ····················· | ···················· | ···················· | ···················· |
| Q | 5577 hCoV-19/France/ARA-SC170/2020\|EPI_ISL_634830\|2020- | ···················· | ···················· | ····················· | ···················· | ···················· | ···················· |
| Q | 5578 hCoV-19/France/ARA-SC173/2020\|EPI_ISL_634831\|2020- | ···················· | ···················· | ····················· | ···················· | ···················· | ···················· |
| Q | 5579 hCoV-19/France/ARA-SC176/2020\|EPI_ISL_634832\|2020- | ···················· | ···················· | ····················· | ···················· | ···················· | ···················· |
| Q | 5580 hCoV-19/France/ARA-SC179/2020\|EPI_ISL_634833\|2020- | ···················· | ···················· | ····················· | ···················· | ···················· | ···················· |
| Q | 5581 hCoV-19/France/ARA-SC180/2020\|EPI_ISL_634834\|2020- | ···················· | ··········A········· | ····················· | ···················· | ···················· | ···················· |
| Q | 5582 hCoV-19/France/ARA-SC183/2020\|EPI_ISL_634835\|2020- | ···················· | ···················· | ····················· | ···················· | ···················· | ···················· |
| Q | 5584 hCoV-19/France/ARA-SC192/2020\|EPI_ISL_634837\|2020- | ···················· | ···················· | ····················· | ···················· | ···················· | ···················· |
| Q | 5585 hCoV-19/USA/MN-MDH-1964/2020\|EPI_ISL_634838\|2020-1 | ···················· | ·A·················· | ····················· | ···················· | ···················· | ···················· |
| Q | 5586 hCoV-19/USA/MN-MDH-1965/2020\|EPI_ISL_634839\|2020-1 | ···················· | ·············A······ | ····················· | ···················· | ···················· | ···················· |
| Q | 5587 hCoV-19/USA/MN-MDH-1966/2020\|EPI_ISL_634840\|2020-1 | ···················· | ·············A······ | ····················· | ···················· | ···················· | ···················· |

Q 5588 hCoV-19/USA/MN-MDH-1967/2020|EPI_ISL_634841|2020-1 ···················· ·············A······ ···················· ···················· ··N················· ····················
Q 5589 hCoV-19/USA/MN-MDH-1968/2020|EPI_ISL_634842|2020-1 ···················· ·············A······ ···················· ···················· ···················· ····················
Q 5590 hCoV-19/USA/MN-MDH-1969/2020|EPI_ISL_634843|2020-1 ···················· ·············A······ ···················· ···················· ···················· ····················
Q 5591 hCoV-19/USA/MN-MDH-1970/2020|EPI_ISL_634844|2020-1 ···················· ·············A······ ···················· ···················· ···················· ····················
Q 5592 hCoV-19/USA/MN-MDH-1971/2020|EPI_ISL_634845|2020-1 ···················· ·············A······ ···················· ···················· ···················· ····················
Q 5593 hCoV-19/USA/MN-MDH-1982/2020|EPI_ISL_634856|2020-1 ···················· ···················· ···················· ···················· ···················· ····················
Q 5594 hCoV-19/USA/MN-MDH-1983/2020|EPI_ISL_634857|2020-1 ···················· ···················· ···················· ···················· ···················· ····················
Q 5595 hCoV-19/USA/MN-MDH-1984/2020|EPI_ISL_634858|2020-1 ···················· ·············A······ ···················· ···················· ···················· ····················
Q 5596 hCoV-19/USA/MN-MDH-1985/2020|EPI_ISL_634859|2020-1 ···················· ···················· ···················· ···················· ···················· ····················
Q 5597 hCoV-19/USA/MN-MDH-1986/2020|EPI_ISL_634860|2020-1 ···················· ···················· ···················· ···················· ···················· ····················
Q 5598 hCoV-19/USA/MN-MDH-1987/2020|EPI_ISL_634861|2020-1 ···················· ···················· ···················· ···················· ···················· ····················
Q 5599 hCoV-19/USA/MN-MDH-1988/2020|EPI_ISL_634862|2020-1 ···················· ···················· ···················· ···················· ···················· ····················
Q 5600 hCoV-19/USA/MN-MDH-1989/2020|EPI_ISL_634863|2020-1 ···················· ···················· ···················· ···················· ···················· ····················
Q 5601 hCoV-19/USA/MN-MDH-1990/2020|EPI_ISL_634864|2020-1 ···················· ···················· ···················· ···················· ···················· ····················
Q 5602 hCoV-19/USA/MN-MDH-1991/2020|EPI_ISL_634865|2020-1 ···················· ···················· ···················· ···················· ···················· ····················
Q 5603 hCoV-19/USA/MN-MDH-1992/2020|EPI_ISL_634866|2020-1 ···················· ·············A······ ···················· ···················· ···················· ····················
Q 5604 hCoV-19/USA/MN-MDH-1993/2020|EPI_ISL_634867|2020-1 ···················· ·············A······ ···················· ···················· ···················· ····················
Q 5605 hCoV-19/USA/MN-MDH-1994/2020|EPI_ISL_634868|2020-1 ···················· ·············A······ ···················· ···················· ···················· ····················
Q 5606 hCoV-19/USA/MN-MDH-1995/2020|EPI_ISL_634869|2020-1 ···················· ·············A······ ···················· ···················· ···················· ····················
Q 5607 hCoV-19/USA/MN-MDH-1996/2020|EPI_ISL_634870|2020-1 ···················· ·A·················· ···················· ···················· ···················· ····················
Q 5608 hCoV-19/USA/MN-MDH-1997/2020|EPI_ISL_634871|2020-1 ···················· ·············A······ ···················· ···················· ···················· ····················
Q 5609 hCoV-19/USA/MN-MDH-1998/2020|EPI_ISL_634872|2020-1 ···················· ·············A······ ···················· ···················· ·NNN················ ···················· 1
Q 5610 hCoV-19/USA/MN-MDH-1999/2020|EPI_ISL_634873|2020-1 ···················· ···················· ···················· ···················· ···················· ····················
Q 5611 hCoV-19/USA/MN-MDH-2000/2020|EPI_ISL_634874|2020-1 ···················· ···················· ···················· ···················· ···················· ····················
Q 5612 hCoV-19/USA/MN-MDH-2001/2020|EPI_ISL_634875|2020-1 ···················· ···················· ···················· ···················· ···················· ····················
Q 5614 hCoV-19/Belgium/UGent-158/2020|EPI_ISL_634879|2020 ···················· ···················· ···················· ···················· ···················· ····················
Q 5616 hCoV-19/Belgium/UGent-164/2020|EPI_ISL_634881|2020 ···················· ···················· ···················· ···················· ···················· ····················
Q 5617 hCoV-19/Belgium/UGent-165/2020|EPI_ISL_634882|2020 ···················· ···················· ···················· ···················· ···················· ····················
Q 5618 hCoV-19/Belgium/UGent-167/2020|EPI_ISL_634883|2020 ···················· ···················· ···················· ···················· ···················· ····················
Q 5619 hCoV-19/Belgium/UGent-168/2020|EPI_ISL_634884|2020 ···················· ···················· ···················· ···················· ···················· ····················
Q 5620 hCoV-19/Belgium/UGent-169/2020|EPI_ISL_634885|2020 ···················· ···················· ···················· ···················· ···················· ····················
Q 5622 hCoV-19/Belgium/UGent-171/2020|EPI_ISL_634887|2020 ···················· ···················· ···················· ···················· ···················· ····················
Q 5623 hCoV-19/Belgium/UGent-172/2020|EPI_ISL_634888|2020 ···················· ···················· ···················· ···················· ···················· ····················
Q 5624 hCoV-19/Belgium/UGent-173/2020|EPI_ISL_634889|2020 ···················· ···················· ···················· ···················· ···················· ····················
Q 5626 hCoV-19/Belgium/UGent-180/2020|EPI_ISL_634891|2020 ···················· ···················· ···················· ···················· ···················· ····················
Q 5627 hCoV-19/Belgium/UGent-181/2020|EPI_ISL_634892|2020 ···················· ···················· ···················· ···················· ···················· ····················
Q 5628 hCoV-19/Belgium/UGent-182/2020|EPI_ISL_634893|2020 ···················· ···················· ···················· ···················· ···················· ····················
Q 5629 hCoV-19/Belgium/UGent-183/2020|EPI_ISL_634894|2020 ···················· ···················· ···················· ···················· ···················· ····················
Q 5630 hCoV-19/Belgium/UGent-184/2020|EPI_ISL_634895|2020 ···················· ···················· ···················· ···················· ···················· ····················
Q 5631 hCoV-19/USA/UT-UPHL-201123379/2020|EPI_ISL_634896| ···················· ···················· ···················· ···················· ···················· ····················
Q 5632 hCoV-19/USA/UT-UPHL-201121906/2020|EPI_ISL_634898| ···················· ···················· ···················· ···················· ···················· ····················
Q 5633 hCoV-19/New Zealand/20CV0513/2020|EPI_ISL_626352|2 ···················· ···················· ···················· ···················· ···················· ····················
Q 5634 hCoV-19/New Zealand/20CV0515/2020|EPI_ISL_626353|2 ···················· ···················· ···················· ···················· ···················· ····················
Q 5635 hCoV-19/New Zealand/20CV0516/2020|EPI_ISL_626354|2 ···················· ···················· ···················· ···················· ···················· ····················
Q 5636 hCoV-19/New Zealand/20CV0517/2020|EPI_ISL_626355|2 ·T·················· ···················· ···················· ···················· ···················· ····················
Q 5637 hCoV-19/New Zealand/20CV0521/2020|EPI_ISL_626356|2 ···················· ·A·················· ···················· ···················· ···················· ··A·················
Q 5638 hCoV-19/USA/IL-NM-0039/2020|EPI_ISL_626359|2020-03 ···················· ···················· ···················· ···················· ···················· ····················
Q 5639 hCoV-19/USA/IL-NM-0052/2020|EPI_ISL_626360|2020-03 ···················· ···················· ···················· ···················· ···················· ····················
Q 5640 hCoV-19/USA/IL-NM-0135/2020|EPI_ISL_626364|2020-03 ···················· ···················· ···················· ···················· ···················· ····················
Q 5641 hCoV-19/USA/IL-NM-0137/2020|EPI_ISL_626365|2020-03 ···················· ···················· ···················· ···················· ···················· ····················
Q 5642 hCoV-19/USA/IL-NM-0142/2020|EPI_ISL_626366|2020-03 ···················· ···················· ···················· ···················· ···················· ····················
Q 5643 hCoV-19/USA/IL-NM-0144/2020|EPI_ISL_626368|2020-03 ···················· ···················· ···················· ···················· ···················· ····················
Q 5644 hCoV-19/USA/IL-NM-0149/2020|EPI_ISL_626370|2020-03 ···················· ···················· ···················· ···················· ···················· ····················
Q 5645 hCoV-19/USA/IL-NM-0153/2020|EPI_ISL_626371|2020-03 ···················· ···················· ···················· ···················· ···················· ····················
Q 5646 hCoV-19/USA/IL-NM-0156/2020|EPI_ISL_626372|2020-03 ···················· ···················· ···················· ···················· ···················· ····················
Q 5647 hCoV-19/USA/IL-NM-0158/2020|EPI_ISL_626373|2020-03 ···················· ···················· ···················· ···················· ···················· ····················
Q 5648 hCoV-19/USA/IL-NM-0173/2020|EPI_ISL_626374|2020-03 ···················· ···················· ···················· ···················· ···················· ····················
Q 5649 hCoV-19/USA/IL-NM-0184/2020|EPI_ISL_626375|2020-03 ···················· ···················· ···················· ···················· ···················· ····················
Q 5650 hCoV-19/USA/IL-NM-0247/2020|EPI_ISL_626376|2020-03 ···················· ···················· ···················· ···················· ···················· ····················
Q 5651 hCoV-19/USA/IL-NM-0267/2020|EPI_ISL_626377|2020-03 ···················· ···················· ···················· ···················· ···················· ····················
Q 5652 hCoV-19/USA/IL-NM-0287/2020|EPI_ISL_626378|2020-03 ···················· ···················· ···················· ···················· ···················· ····················
Q 5653 hCoV-19/USA/IL-NM-0329/2020|EPI_ISL_626379|2020-03 ···················· ···················· ···················· ···················· ···················· ····················
Q 5654 hCoV-19/USA/IL-NM-0353/2020|EPI_ISL_626381|2020-03 ···················· ···················· ···················· ···················· ···················· ····················
Q 5655 hCoV-19/USA/IL-NM-0367/2020|EPI_ISL_626382|2020-03 ···················· ···················· ···················· ···················· ···················· ····················
Q 5656 hCoV-19/USA/IL-NM-0376/2020|EPI_ISL_626383|2020-03 ···················· ···················· ···················· ···················· ···················· ····················
Q 5657 hCoV-19/USA/IL-NM-0382/2020|EPI_ISL_626384|2020-03 ···················· ···················· ···················· ···················· ···················· ····················
Q 5658 hCoV-19/USA/IL-NM-0395/2020|EPI_ISL_626385|2020-04 ···················· ···················· ···················· ···················· ···················· ····················
Q 5659 hCoV-19/USA/IL-NM-0396/2020|EPI_ISL_626386|2020-04 ···················· ···················· ···················· ···················· ···················· ····················
Q 5660 hCoV-19/USA/IL-NM-0397/2020|EPI_ISL_626387|2020-04 ···················· ···················· ···················· ···················· ···················· ····················
Q 5661 hCoV-19/USA/IL-NM-0398/2020|EPI_ISL_626388|2020-04 ···················· ···················· ···················· ···················· ···················· ····················
Q 5662 hCoV-19/USA/IL-NM-0399/2020|EPI_ISL_626389|2020-04 ···················· ···················· ···················· ···················· ···················· ····················
Q 5663 hCoV-19/USA/IL-NM-0404/2020|EPI_ISL_626390|2020-04 ···················· ···················· ···················· ···················· ···················· ····················
Q 5664 hCoV-19/USA/IL-NM-0405/2020|EPI_ISL_626391|2020-04 ···················· ···················· ···················· ···················· ···················· ····················
Q 5665 hCoV-19/USA/IL-NM-0410/2020|EPI_ISL_626392|2020-04 ···················· ···················· ···················· ···················· ···················· ····················
Q 5666 hCoV-19/USA/IL-NM-0411/2020|EPI_ISL_626393|2020-04 ···················· ···················· ···················· ···················· ···················· ····················
Q 5667 hCoV-19/USA/IL-NM-0413/2020|EPI_ISL_626394|2020-04 ···················· ···················· ···················· ···················· ···················· ····················
Q 5668 hCoV-19/USA/IL-NM-0414/2020|EPI_ISL_626395|2020-04 ···················· ···················· ···················· ···················· ···················· ····················
Q 5669 hCoV-19/USA/IL-NM-0415/2020|EPI_ISL_626396|2020-04 ···················· ···················· ···················· ···················· ···················· ····················
Q 5670 hCoV-19/USA/IL-NM-0416/2020|EPI_ISL_626397|2020-04 ···················· ···················· ···················· ···················· ···················· ····················
Q 5671 hCoV-19/USA/IL-NM-0417/2020|EPI_ISL_626398|2020-04 ···················· ···················· ···················· ···················· ···················· ····················

| | | | | | | | |
|---|---|---|---|---|---|---|---|
| Q | 5672 hCoV-19/USA/IL-NM-0419/2020\|EPI_ISL_626399\|2020-04 | ···················· | ···················· | ····················· | ···················· | ···················· | ···················· |
| Q | 5673 hCoV-19/England/NORW-F1503/2020\|EPI_ISL_627600\|202 | ···················· | ···················· | ····················· | ···················· | ···················· | ···················· |
| Q | 5675 hCoV-19/England/NORW-F1530/2020\|EPI_ISL_627602\|202 | ···················· | ···················· | ····················· | ···················· | ···················· | ···················· |
| Q | 5676 hCoV-19/England/NORW-F155E/2020\|EPI_ISL_627603\|202 | ···················· | ···················· | ····················· | ···················· | ···················· | ···················· |
| Q | 5678 hCoV-19/England/NORW-F158B/2020\|EPI_ISL_627605\|202 | ···················· | ···················· | ····················· | ···················· | ···················· | ···················· |
| Q | 5679 hCoV-19/England/NORW-F15A9/2020\|EPI_ISL_627606\|202 | ···················· | ···················· | ····················· | ···················· | ···················· | ···················· |
| Q | 5680 hCoV-19/England/NORW-F15C7/2020\|EPI_ISL_627607\|202 | ···················· | ···················· | ····················· | ···················· | ···················· | ···················· |
| Q | 5682 hCoV-19/England/NORW-F163D/2020\|EPI_ISL_627609\|202 | ···················· | ···················· | ····················· | ···················· | ···················· | ···················· |
| Q | 5683 hCoV-19/England/NORW-F164C/2020\|EPI_ISL_627610\|202 | ···················· | ···················· | ····················· | ···················· | ···················· | ···················· |
| Q | 5685 hCoV-19/England/NORW-F1697/2020\|EPI_ISL_627612\|202 | ···················· | ···················· | ····················· | ···················· | ···················· | ···················· |
| Q | 5687 hCoV-19/England/NORW-F170D/2020\|EPI_ISL_627614\|202 | ···················· | ···················· | ····················· | ···················· | ···················· | ···················· |
| Q | 5690 hCoV-19/England/NORW-F17EF/2020\|EPI_ISL_627617\|202 | ···················· | ···················· | ····················· | ···················· | ···················· | ···················· |
| Q | 5691 hCoV-19/England/NORW-F1B2F/2020\|EPI_ISL_627618\|202 | ···················· | ···················· | ····················· | ···················· | ···················· | ···················· |
| Q | 5692 hCoV-19/England/NORW-F1B6B/2020\|EPI_ISL_627619\|202 | ···················· | ·A·················· | ····················· | ···················· | ···················· | ··A················· |
| Q | 5693 hCoV-19/England/NORW-F1BA7/2020\|EPI_ISL_627620\|202 | ···················· | ···················· | ····················· | ···················· | ···················· | ···················· |
| Q | 5694 hCoV-19/England/NORW-F1BD4/2020\|EPI_ISL_627621\|202 | ···················· | ···················· | ····················· | ···················· | ···················· | ···················· |
| Q | 5695 hCoV-19/England/NORW-F1BE3/2020\|EPI_ISL_627622\|202 | ···················· | ···················· | ····················· | ···················· | ···················· | ···················· |
| Q | 5696 hCoV-19/England/NORW-F1C0E/2020\|EPI_ISL_627623\|202 | ···················· | ···················· | ····················· | ···················· | ···················· | ···················· |
| Q | 5697 hCoV-19/England/NORW-F1C1D/2020\|EPI_ISL_627624\|202 | ···················· | ···················· | ····················· | ···················· | ···················· | ···················· |
| Q | 5698 hCoV-19/England/NORW-F1C2C/2020\|EPI_ISL_627625\|202 | ···················· | ···················· | ····················· | ···················· | ···················· | ···················· |
| Q | 5699 hCoV-19/England/NORW-F1C68/2020\|EPI_ISL_627626\|202 | ···················· | ···················· | ····················· | ···················· | ···················· | ···················· |
| Q | 5700 hCoV-19/England/NORW-F1C95/2020\|EPI_ISL_627627\|202 | ···················· | ···················· | ····················· | ···················· | ···················· | ···················· |
| Q | 5701 hCoV-19/England/NORW-F1CA4/2020\|EPI_ISL_627628\|202 | ···················· | ···················· | ····················· | ···················· | ···················· | ···················· |
| Q | 5702 hCoV-19/England/NORW-F1CB3/2020\|EPI_ISL_627629\|202 | ···················· | ···················· | ····················· | ···················· | ···················· | ···················· |
| Q | 5704 hCoV-19/England/NORW-F1CE0/2020\|EPI_ISL_627631\|202 | ···················· | ···················· | ····················· | ···················· | ···················· | N··················· |
| Q | 5705 hCoV-19/England/NORW-F1D0B/2020\|EPI_ISL_627632\|202 | ···················· | ···················· | ····················· | ···················· | ···················· | ···················· |
| Q | 5706 hCoV-19/England/NORW-F1D47/2020\|EPI_ISL_627633\|202 | ···················· | ···················· | ····················· | ···················· | ···················· | ···················· |
| Q | 5708 hCoV-19/England/NORW-F1D74/2020\|EPI_ISL_627635\|202 | ···················· | ···················· | ····················· | ···················· | ···················· | ···················· |
| Q | 5709 hCoV-19/England/NORW-F1DA1/2020\|EPI_ISL_627636\|202 | ···················· | ···················· | ····················· | ···················· | ···················· | ···················· |
| Q | 5710 hCoV-19/England/NORW-F1DCF/2020\|EPI_ISL_627637\|202 | ···················· | ···················· | ····················· | ···················· | ···················· | ···················· |
| Q | 5711 hCoV-19/England/NORW-F1DED/2020\|EPI_ISL_627638\|202 | ···················· | ···················· | ····················· | ···················· | ···················· | ···················· |
| Q | 5713 hCoV-19/England/NORW-F1E26/2020\|EPI_ISL_627640\|202 | ···················· | ···················· | ····················· | ···················· | ···················· | ···················· |
| Q | 5714 hCoV-19/England/NORW-F1E35/2020\|EPI_ISL_627641\|202 | ···················· | ···················· | ····················· | ···················· | ···················· | ···················· |
| Q | 5716 hCoV-19/England/NORW-F1F14/2020\|EPI_ISL_627643\|202 | ···················· | ···················· | ····················· | ···················· | ···················· | ···················· |
| Q | 5718 hCoV-19/England/NORW-F1FAB/2020\|EPI_ISL_627645\|202 | ···················· | ···················· | ····················· | ···················· | ···················· | ···················· |
| Q | 5720 hCoV-19/England/NORW-F1FD8/2020\|EPI_ISL_627647\|202 | ···················· | ···················· | ····················· | ···················· | ···················· | ···················· |
| Q | 5721 hCoV-19/England/NORW-F1FE7/2020\|EPI_ISL_627648\|202 | ···················· | ···················· | ····················· | ···················· | ···················· | ···················· |
| Q | 5722 hCoV-19/England/NORW-F2001/2020\|EPI_ISL_627649\|202 | ···················· | ···················· | ····················· | ···················· | ···················· | ···················· |
| Q | 5723 hCoV-19/England/NORW-F202F/2020\|EPI_ISL_627650\|202 | ···················· | ···················· | ····················· | ···················· | ···················· | ···················· |
| Q | 5724 hCoV-19/England/NORW-F204D/2020\|EPI_ISL_627651\|202 | ···················· | ···················· | ····················· | ···················· | ···················· | ···················· |
| Q | 5725 hCoV-19/England/NORW-F206B/2020\|EPI_ISL_627652\|202 | ···················· | ···················· | ····················· | ···················· | ···················· | ···················· |
| Q | 5726 hCoV-19/England/NORW-F207A/2020\|EPI_ISL_627653\|202 | ···················· | ···················· | ····················· | ···················· | ···················· | ···················· |
| Q | 5727 hCoV-19/England/NORW-F20B6/2020\|EPI_ISL_627654\|202 | ···················· | ···················· | ····················· | ···················· | ···················· | ···················· |
| Q | 5728 hCoV-19/England/NORW-F213B/2020\|EPI_ISL_627655\|202 | ···················· | ···················· | ····················· | ···················· | ···················· | ···················· |
| Q | 5729 hCoV-19/England/NORW-F2159/2020\|EPI_ISL_627656\|202 | ···················· | ···················· | ····················· | ···················· | ···················· | ···················· |
| Q | 5730 hCoV-19/England/NORW-F2168/2020\|EPI_ISL_627657\|202 | ···················· | ···················· | ····················· | ···················· | ···················· | ···················· |
| Q | 5731 hCoV-19/England/NORW-F2186/2020\|EPI_ISL_627658\|202 | ···················· | ···················· | ····················· | ···················· | ···················· | ···················· |
| Q | 5733 hCoV-19/England/NORW-F2265/2020\|EPI_ISL_627660\|202 | ···················· | ···················· | ····················· | ···················· | ···················· | ···················· |
| Q | 5736 hCoV-19/England/NORW-F23CC/2020\|EPI_ISL_627663\|202 | ···················· | ···················· | ····················· | ···················· | ···················· | ···················· |
| Q | 5737 hCoV-19/England/NORW-F3CB1/2020\|EPI_ISL_627664\|202 | ···················· | ···················· | ····················· | ···················· | ···················· | ···················· |
| Q | 5738 hCoV-19/England/NORW-F3E15/2020\|EPI_ISL_627665\|202 | ···················· | ···················· | ····················· | ···················· | ···················· | ···················· |
| Q | 5739 hCoV-19/England/NOTT-116A80/2020\|EPI_ISL_627666\|20 | ···················· | ···················· | ····················· | ···················· | ···················· | ···················· |
| Q | 5740 hCoV-19/England/NOTT-116D96/2020\|EPI_ISL_627667\|20 | ···················· | ···················· | ····················· | ···················· | ···················· | ···················· |
| Q | 5741 hCoV-19/England/NOTT-116DA5/2020\|EPI_ISL_627668\|20 | ···················· | ···················· | ····················· | ···················· | ···················· | ···················· |
| Q | 5742 hCoV-19/England/NOTT-117032/2020\|EPI_ISL_627669\|20 | ···················· | ···················· | ····················· | ···················· | ···················· | ···················· |
| Q | 5743 hCoV-19/England/NOTT-1175F7/2020\|EPI_ISL_627670\|20 | ···················· | ···················· | ····················· | ···················· | ···················· | ···················· |
| Q | 5744 hCoV-19/England/NOTT-117876/2020\|EPI_ISL_627671\|20 | ···················· | ···················· | ····················· | ···················· | ···················· | ···················· |
| Q | 5745 hCoV-19/England/NOTT-117B8C/2020\|EPI_ISL_627672\|20 | ···················· | ···················· | ····················· | ···················· | ···················· | ···················· |
| Q | 5746 hCoV-19/England/NOTT-117E38/2020\|EPI_ISL_627673\|20 | ···················· | ···················· | ····················· | ···················· | ···················· | ···················· |
| Q | 5747 hCoV-19/England/NOTT-11831A/2020\|EPI_ISL_627674\|20 | ···················· | ···················· | ····················· | ···················· | ···················· | ···················· |
| Q | 5748 hCoV-19/England/NOTT-1184DB/2020\|EPI_ISL_627675\|20 | ···················· | ···················· | ····················· | ···················· | ···················· | ···················· |
| Q | 5749 hCoV-19/England/NOTT-11857E/2020\|EPI_ISL_627676\|20 | ···················· | ···················· | ····················· | ···················· | ···················· | ···················· |
| Q | 5750 hCoV-19/England/NOTT-118620/2020\|EPI_ISL_627677\|20 | ···················· | ···················· | ····················· | ···················· | ···················· | ···················· |
| Q | 5751 hCoV-19/England/NOTT-11863F/2020\|EPI_ISL_627678\|20 | ···················· | ···················· | ····················· | ···················· | ···················· | ···················· |
| Q | 5752 hCoV-19/England/NOTT-11865D/2020\|EPI_ISL_627679\|20 | ···················· | ···················· | ····················· | ···················· | ···················· | ···················· |
| Q | 5753 hCoV-19/England/NOTT-118699/2020\|EPI_ISL_627680\|20 | ···················· | ···················· | ····················· | ···················· | ···················· | ···················· |
| Q | 5754 hCoV-19/England/NOTT-1186A8/2020\|EPI_ISL_627681\|20 | ···················· | ···················· | ····················· | ···················· | ···················· | ···················· |
| Q | 5755 hCoV-19/England/NOTT-1186B7/2020\|EPI_ISL_627682\|20 | ···················· | ···················· | ····················· | ···················· | ···················· | ···················· |
| Q | 5756 hCoV-19/England/NOTT-118778/2020\|EPI_ISL_627683\|20 | ···················· | ···················· | ····················· | ···················· | ···················· | ···················· |
| Q | 5757 hCoV-19/England/NOTT-11881B/2020\|EPI_ISL_627684\|20 | ···················· | ···················· | ····················· | ···················· | ···················· | ···················· |
| Q | 5758 hCoV-19/England/NOTT-118945/2020\|EPI_ISL_627685\|20 | ···················· | ···················· | ····················· | ···················· | ···················· | ···················· |
| Q | 5759 hCoV-19/England/NOTT-118990/2020\|EPI_ISL_627686\|20 | ···················· | ···················· | ····················· | ···················· | ···················· | ···················· |
| Q | 5760 hCoV-19/England/NOTT-118A7F/2020\|EPI_ISL_627687\|20 | ···················· | ···················· | ····················· | ···················· | ···················· | ···················· |
| Q | 5761 hCoV-19/England/NOTT-118A8E/2020\|EPI_ISL_627688\|20 | ···················· | ···················· | ····················· | ···················· | ···················· | ···················· |
| Q | 5762 hCoV-19/England/NOTT-118A9D/2020\|EPI_ISL_627689\|20 | ···················· | ···················· | ····················· | ···················· | ···················· | ···················· |
| Q | 5763 hCoV-19/England/NOTT-118AAC/2020\|EPI_ISL_627690\|20 | ···················· | ···················· | ····················· | ···················· | ···················· | ···················· |
| Q | 5764 hCoV-19/England/NOTT-118ABB/2020\|EPI_ISL_627691\|20 | ···················· | ···················· | ····················· | ···················· | ···················· | ···················· |
| Q | 5765 hCoV-19/England/NOTT-118C79/2020\|EPI_ISL_627692\|20 | ···················· | ···················· | ····················· | ···················· | ···················· | ···················· |
| Q | 5766 hCoV-19/England/NOTT-118D3A/2020\|EPI_ISL_627693\|20 | ···················· | ···················· | ····················· | ···················· | ···················· | ···················· |
| Q | 5767 hCoV-19/England/NOTT-118D58/2020\|EPI_ISL_627694\|20 | ···················· | ···················· | ····················· | ···················· | ···················· | ···················· |

```
Q 5768 hCoV-19/England/NOTT-118D76/2020|EPI_ISL_627695|20 .................... .................... .................... .................... .................... ....................
Q 5769 hCoV-19/England/NOTT-118E28/2020|EPI_ISL_627696|20 .................... .................... .................... .................... .................... ....................
Q 5770 hCoV-19/England/NOTT-118E55/2020|EPI_ISL_627697|20 .................... .................... .................... .................... .................... ....................
Q 5771 hCoV-19/England/NOTT-118E73/2020|EPI_ISL_627698|20 .................... .................... .................... .................... .................... ....................
Q 5772 hCoV-19/England/NOTT-118E82/2020|EPI_ISL_627699|20 .................... .................... .................... .................... .................... ....................
Q 5773 hCoV-19/USA/UT-UPHL-201113771/2020|EPI_ISL_628932| .................... .A.................. .................... .................... .................... ....................
Q 5774 hCoV-19/USA/UT-UPHL-201102369/2020|EPI_ISL_628933| .................... .................... .................... .................... .................... ....................
Q 5775 hCoV-19/USA/UT-UPHL-201131911/2020|EPI_ISL_628934| .................... .................... .................... .................... .................... ....................
Q 5776 hCoV-19/USA/UT-UPHL-201110826/2020|EPI_ISL_628935| .................... .................... .................... .................... .................... ....................
Q 5777 hCoV-19/USA/UT-UPHL-201126070/2020|EPI_ISL_628936| .................... .................... .................... .................... .................... ....................
Q 5778 hCoV-19/USA/UT-UPHL-201109633/2020|EPI_ISL_628937| .................... .................... .................... .................... .................... ....................
Q 5779 hCoV-19/USA/UT-UPHL-201110857/2020|EPI_ISL_628938| .................... .A.................. .................... .................... .................... ....................
Q 5780 hCoV-19/USA/UT-UPHL-201116688/2020|EPI_ISL_628939| .................... .................... .................... .................... .................... ....................
Q 5781 hCoV-19/USA/UT-UPHL-201125055/2020|EPI_ISL_628940| .................... .................... .................... .................... .................... ....................
Q 5782 hCoV-19/USA/UT-UPHL-201119442/2020|EPI_ISL_628941| .................... .................... .................... .................... .................... ....................
Q 5783 hCoV-19/USA/UT-UPHL-201103044/2020|EPI_ISL_628943| .................... .................... .................... .................... .................... ....................
Q 5784 hCoV-19/USA/UT-UPHL-201128848/2020|EPI_ISL_628945| .................... .................... .................... .................... .................... ....................
Q 5785 hCoV-19/USA/UT-UPHL-201108217/2020|EPI_ISL_628951| .................... .................... .................... .................... .................... ....................
Q 5786 hCoV-19/USA/UT-UPHL-201103500/2020|EPI_ISL_628953| .................... .A.................. .................... .................... .................... ....................
Q 5787 hCoV-19/USA/UT-UPHL-201124588/2020|EPI_ISL_628954| .................... .A.................. .................... .................... .................... ....................
Q 5788 hCoV-19/USA/UT-UPHL-201101052/2020|EPI_ISL_628955| .................... .A.................. .................... .................... .................... ....................
Q 5789 hCoV-19/USA/UT-UPHL-201106283/2020|EPI_ISL_628958| .................... .................... .................... .................... .................... ....................
Q 5790 hCoV-19/USA/UT-UPHL-201108594/2020|EPI_ISL_628959| .................... .................... .................... .................... .................... ....................
Q 5791 hCoV-19/USA/UT-UPHL-201117948/2020|EPI_ISL_628960| .................... .................... .................... .................... .................... ....................
Q 5792 hCoV-19/USA/UT-UPHL-201114272/2020|EPI_ISL_628961| .................... .A.................. .................... .................... .................... ....................
Q 5793 hCoV-19/USA/UT-UPHL-201122169/2020|EPI_ISL_628964| .................... .................... .................... .................... .................... ....................
Q 5794 hCoV-19/USA/UT-UPHL-201122286/2020|EPI_ISL_628967| .................... .................... .................... .................... .................... ....................
Q 5795 hCoV-19/USA/UT-UPHL-201105786/2020|EPI_ISL_628968| .................... .................... .................... .................... .................... ....................
Q 5796 hCoV-19/USA/UT-UPHL-201118881/2020|EPI_ISL_628969| .................... .................... .................... .................... .................... ....................
Q 5797 hCoV-19/USA/UT-UPHL-201132584/2020|EPI_ISL_628971| .................... .................... .................... .................... .................... ....................
Q 5798 hCoV-19/USA/UT-UPHL-201101361/2020|EPI_ISL_628972| .................... .................... .................... .................... .................... ....................
Q 5799 hCoV-19/USA/UT-UPHL-201103306/2020|EPI_ISL_628975| .................... .................... .................... .................... .................... ....................
Q 5800 hCoV-19/USA/UT-UPHL-201126930/2020|EPI_ISL_628976| .................... .................... .................... .................... .................... ....................
Q 5801 hCoV-19/USA/UT-UPHL-201123424/2020|EPI_ISL_628977| .................... .................... .................... .................... .................... ....................
Q 5802 hCoV-19/USA/UT-UPHL-201116313/2020|EPI_ISL_628978| .................... .................... .................... .................... .........A.......... ....................
Q 5803 hCoV-19/USA/UT-UPHL-201121140/2020|EPI_ISL_628979| .................... .................... .................... .................... .................... ....................
Q 5804 hCoV-19/USA/UT-UPHL-201101119/2020|EPI_ISL_628981| .................... .................... .................... .................... .................... ....................
Q 5805 hCoV-19/USA/UT-UPHL-201108243/2020|EPI_ISL_628983| .................... .................... .................... .................... .................... ....................
Q 5806 hCoV-19/USA/UT-UPHL-201109408/2020|EPI_ISL_628984| .................... .................... .................... .................... .................... ....................
Q 5807 hCoV-19/USA/UT-UPHL-201115941/2020|EPI_ISL_628985| .................... .................... .................... .................... .................... ....................
Q 5808 hCoV-19/USA/UT-UPHL-201110199/2020|EPI_ISL_628986| .................... .................... .................... .................... .................... ....................
Q 5809 hCoV-19/USA/UT-UPHL-201132476/2020|EPI_ISL_628987| .................... .................... .................... .................... .................... ....................
Q 5810 hCoV-19/USA/UT-UPHL-201101347/2020|EPI_ISL_628989| .................... .................... .................... .................... .................... ....................
Q 5811 hCoV-19/USA/UT-UPHL-201117148/2020|EPI_ISL_628990| .................... .................... .................... .................... .................... ....................
Q 5812 hCoV-19/USA/UT-UPHL-201124734/2020|EPI_ISL_628991| .................... .................... .................... .................... .................... ....................
Q 5813 hCoV-19/USA/UT-UPHL-201131401/2020|EPI_ISL_628992| .................... .................... .................... .................... .................... ....................
Q 5814 hCoV-19/USA/UT-UPHL-201114411/2020|EPI_ISL_628993| .................... .................... .................... .................... .................... ....................
Q 5815 hCoV-19/USA/UT-UPHL-201114492/2020|EPI_ISL_628994| .................... .................... .................... .................... .................... ....................
Q 5816 hCoV-19/USA/UT-UPHL-201118192/2020|EPI_ISL_628996| .................... .................... .................... .................... .................... ....................
Q 5817 hCoV-19/USA/UT-UPHL-201121026/2020|EPI_ISL_628997| .................... .................... .................... .................... .................... ....................
Q 5818 hCoV-19/USA/UT-UPHL-201113918/2020|EPI_ISL_628998| .................... .................... .................... .................... .................... ....................
Q 5819 hCoV-19/USA/UT-UPHL-201131364/2020|EPI_ISL_628999| .................... .................... .................... .................... .................... ....................
Q 5820 hCoV-19/USA/NY-NYCPHL-000800/2020|EPI_ISL_632180|2 .................... .................... .................... .................... .................... ....................
Q 5821 hCoV-19/USA/NY-NYCPHL-000801/2020|EPI_ISL_632181|2 .................... .................... .................... .................... .................... ....................
Q 5822 hCoV-19/USA/NY-NYCPHL-000802/2020|EPI_ISL_632182|2 .................... .................... .................... .................... .................... ....................
Q 5823 hCoV-19/USA/NY-NYCPHL-000803/2020|EPI_ISL_632183|2 .................... .................... .................... .................... .................... ....................
Q 5824 hCoV-19/USA/NY-NYCPHL-000804/2020|EPI_ISL_632184|2 .................... .................... .................... .................... .................... ....................
Q 5825 hCoV-19/USA/NY-NYCPHL-000806/2020|EPI_ISL_632185|2 .................... .................... .................... .................... .................... ....................
Q 5826 hCoV-19/USA/NY-NYCPHL-000807/2020|EPI_ISL_632186|2 .................... .................... .................... .................... .................... ....................
Q 5827 hCoV-19/USA/NY-NYCPHL-000808/2020|EPI_ISL_632187|2 .................... .................... .................... .................... .................... ....................
Q 5828 hCoV-19/USA/NY-NYCPHL-000809/2020|EPI_ISL_632188|2 .................... .................... .................... .................... .................... ....................
Q 5829 hCoV-19/USA/NY-NYCPHL-000810/2020|EPI_ISL_632189|2 .................... .................... .................... .................... .................... ....................
Q 5830 hCoV-19/USA/NY-NYCPHL-000811/2020|EPI_ISL_632190|2 .................... .................... .................... .................... .................... ....................
Q 5831 hCoV-19/USA/NY-NYCPHL-000813/2020|EPI_ISL_632191|2 .................... .................... .................... .................... .................... ....................
Q 5832 hCoV-19/USA/NY-NYCPHL-000815/2020|EPI_ISL_632192|2 .................... .................... .................... .................... .................... ....................
Q 5833 hCoV-19/USA/NY-NYCPHL-000816/2020|EPI_ISL_632193|2 .................... .................... .................... .................... .................... ....................
Q 5834 hCoV-19/USA/NY-NYCPHL-000817/2020|EPI_ISL_632194|2 .................... .................... .................... .................... .................... ....................
Q 5835 hCoV-19/USA/NY-NYCPHL-000818/2020|EPI_ISL_632195|2 .................... .................... .................... .................... .................... ....................
Q 5836 hCoV-19/USA/NY-NYCPHL-000819/2020|EPI_ISL_632196|2 .................... .................... .................... .................... .................... ....................
Q 5837 hCoV-19/USA/NY-NYCPHL-000820/2020|EPI_ISL_632197|2 .................... .................... .................... .................... .................... ....................
Q 5838 hCoV-19/USA/NY-NYCPHL-000821/2020|EPI_ISL_632198|2 .................... .................... .................... .................... .................... ....................
Q 5839 hCoV-19/USA/NY-NYCPHL-000822/2020|EPI_ISL_632199|2 .................... .................... .................... .................... .................... ....................
Q 5840 hCoV-19/USA/NY-NYCPHL-000862/2020|EPI_ISL_632238|2 .................... .................... .................... .................... .................... ....................
Q 5841 hCoV-19/USA/NY-NYCPHL-000863/2020|EPI_ISL_632239|2 .................... .................... .................... .................... .................... ....................
Q 5842 hCoV-19/USA/NY-NYCPHL-000865/2020|EPI_ISL_632241|2 .................... .................... .................... .................... .................... ....................
Q 5843 hCoV-19/Scotland/GCVR-16FC46/2020|EPI_ISL_632242|2 .................... .................... .................... .................... .................... ....................
Q 5844 hCoV-19/England/QEUH-AE4E95/2020|EPI_ISL_633400|20 .................... .................... .................... .................... .................... ....................
Q 5845 hCoV-19/England/QEUH-AE4D7A/2020|EPI_ISL_633401|20 .................... .................... .................... .................... .................... ...A................
Q 5847 hCoV-19/England/CAMC-AEA72B/2020|EPI_ISL_633403|20 .................... .................... .................... .................... .................... ....................
Q 5848 hCoV-19/England/QEUH-AE2705/2020|EPI_ISL_633415|20 .................... .................... .................... .................... .................... ....................
```

Q 5849 hCoV-19/England/QEUH-AE1EB6/2020|EPI_ISL_633417|20 ···················· ···················· ····················· ···················· ···················· ····················
Q 5850 hCoV-19/England/QEUH-AE2D9A/2020|EPI_ISL_633418|20 ···················· ···················· ····················· ···················· ···················· ····················
Q 5851 hCoV-19/England/QEUH-AE2671/2020|EPI_ISL_633419|20 ···················· ···················· ····················· ···················· ···················· ····················
Q 5852 hCoV-19/England/QEUH-AE2565/2020|EPI_ISL_633420|20 ···················· ···················· ····················· ···················· ···················· ····················
Q 5853 hCoV-19/England/QEUH-AE318E/2020|EPI_ISL_633422|20 ···················· ···················· ····················· ···················· ···················· ····················
Q 5854 hCoV-19/England/QEUH-AE229B/2020|EPI_ISL_633423|20 ···················· ···················· ····················· ···················· ···················· ····················
Q 5855 hCoV-19/England/QEUH-AE2AA2/2020|EPI_ISL_633424|20 ···················· ···················· ····················· ···················· ···················· ····················
Q 5856 hCoV-19/England/QEUH-AE2A57/2020|EPI_ISL_633425|20 ···················· ···················· ····················· ···················· ···················· ····················
Q 5857 hCoV-19/England/QEUH-AE1DF5/2020|EPI_ISL_633572|20 ···················· ···················· ····················· ···················· ···················· ····················
Q 5858 hCoV-19/England/QEUH-AE25B0/2020|EPI_ISL_633573|20 ···················· ···················· ····················· ···················· ···················· ····················
Q 5859 hCoV-19/England/QEUH-AE2B18/2020|EPI_ISL_633574|20 ···················· ···················· ····················· ···················· ···················· ····················
Q 5860 hCoV-19/England/QEUH-AE1AC1/2020|EPI_ISL_633575|20 ···················· ···················· ····················· ···················· ···················· ····················
Q 5861 hCoV-19/England/QEUH-AE3036/2020|EPI_ISL_633576|20 ···················· ···················· ····················· ···················· ···················· ····················
Q 5862 hCoV-19/England/QEUH-AD419F/2020|EPI_ISL_634714|20 ···················· ···················· ····················· ···················· ···················· ····················
Q 5863 hCoV-19/England/QEUH-AD56EA/2020|EPI_ISL_634715|20 ···················· ···················· ····················· ···················· ···················· ····················
Q 5864 hCoV-19/England/QEUH-AD400B/2020|EPI_ISL_634716|20 ···················· ···················· ····················· ···················· ···················· ····················
Q 5865 hCoV-19/England/QEUH-AD583F/2020|EPI_ISL_634717|20 ···················· ···················· ····················· ···················· ···················· ····················
Q 5866 hCoV-19/England/QEUH-AD579C/2020|EPI_ISL_634718|20 ···················· ···················· ····················· ···················· ···················· ····················
Q 5867 hCoV-19/England/QEUH-AD103B/2020|EPI_ISL_634720|20 ···················· ···················· ····················· ···················· ···················· ····················
Q 5868 hCoV-19/England/QEUH-AD3F96/2020|EPI_ISL_634721|20 ···················· ···················· ····················· ···················· ···················· ····················
Q 5869 hCoV-19/England/QEUH-AD4672/2020|EPI_ISL_634722|20 ···················· ···················· ····················· ···················· ···················· ····················
Q 5870 hCoV-19/England/QEUH-AD1095/2020|EPI_ISL_634723|20 ···················· ···················· ····················· ···················· ···················· ····················
Q 5871 hCoV-19/England/QEUH-AD1F89/2020|EPI_ISL_634725|20 ···················· ···················· ····················· ···················· ···················· ····················
Q 5872 hCoV-19/England/QEUH-AD19A9/2020|EPI_ISL_634726|20 ···················· ···················· ····················· ···················· ···················· ····················
Q 5873 hCoV-19/England/QEUH-AD5EC4/2020|EPI_ISL_634727|20 ···················· ···················· ····················· ···················· ···················· ····················
Q 5874 hCoV-19/England/QEUH-AD56DB/2020|EPI_ISL_634728|20 ···················· ···················· ····················· ···················· ···················· ····················
Q 5875 hCoV-19/England/QEUH-AD5F1C/2020|EPI_ISL_634729|20 ···················· ···················· ····················· ···················· ···················· ····················
Q 5876 hCoV-19/England/QEUH-AD0FB7/2020|EPI_ISL_634730|20 ···················· ···················· ····················· ···················· ···················· ····················
Q 5877 hCoV-19/England/QEUH-AD4162/2020|EPI_ISL_634731|20 ···················· ···················· ····················· ···················· ···················· ····················
Q 5878 hCoV-19/England/QEUH-AD18CA/2020|EPI_ISL_634732|20 ···················· ···················· ····················· ···················· ···················· ····················
Q 5879 hCoV-19/England/QEUH-AD5F2B/2020|EPI_ISL_634733|20 ···················· ···················· ····················· ···················· ···················· ····················
Q 5880 hCoV-19/Wales/QEUH-AD3E99/2020|EPI_ISL_634748|2020 ···················· ···················· ····················· ···················· ···················· ····················
Q 5881 hCoV-19/Wales/QEUH-AD34D3/2020|EPI_ISL_634749|2020 ···················· ···················· ····················· ···················· ···················· ····················
Q 5882 hCoV-19/Wales/QEUH-AD261A/2020|EPI_ISL_634750|2020 ···················· ···················· ····················· ···················· ···················· ····················
Q 5883 hCoV-19/Wales/QEUH-AD3EB7/2020|EPI_ISL_634751|2020 ···················· ···················· ····················· ···················· ···················· ····················
Q 5884 hCoV-19/Wales/QEUH-AD1781/2020|EPI_ISL_634752|2020 ···················· ···················· ····················· ···················· ···················· ····················
Q 5885 hCoV-19/Wales/QEUH-AD3A59/2020|EPI_ISL_634753|2020 ···················· ···················· ····················· ···················· ···················· ····················
Q 5886 hCoV-19/Wales/ALDP-A675A4/2020|EPI_ISL_634754|2020 ···················· ···················· ····················· ···················· ···················· ····················
Q 5887 hCoV-19/Wales/ALDP-A6754A/2020|EPI_ISL_634755|2020 ···················· ···················· ····················· ···················· ···················· ····················
Q 5888 hCoV-19/Wales/ALDP-A675D1/2020|EPI_ISL_634756|2020 ···················· ···················· ····················· ···················· ···················· ····················
Q 5889 hCoV-19/England/MILK-A6A4B3/2020|EPI_ISL_627508|20 ···················· ···················· ····················· ···················· ···················· ····················
Q 5890 hCoV-19/England/MILK-A6A4C2/2020|EPI_ISL_627509|20 ···················· ···················· ····················· ···················· ···················· ····················
Q 5891 hCoV-19/England/MILK-A6A51A/2020|EPI_ISL_627510|20 ···················· ···················· ····················· ···················· ···················· ····················
Q 5892 hCoV-19/England/MILK-A6A583/2020|EPI_ISL_627511|20 ···················· ···················· ····················· ···················· ···················· ····················
Q 5893 hCoV-19/England/MILK-A6A5A1/2020|EPI_ISL_627512|20 ···················· ···················· ····················· ···················· ···················· ····················
Q 5894 hCoV-19/Scotland/MILK-A6A5CF/2020|EPI_ISL_627513|2 ···················· ···················· ····················· ···················· ···················· ····················
Q 5895 hCoV-19/England/MILK-A6A6BD/2020|EPI_ISL_627514|20 ···················· ···················· ····················· ···················· ···················· ····················
Q 5896 hCoV-19/England/MILK-A6A6DB/2020|EPI_ISL_627515|20 ···················· ···················· ····················· ···················· ······T············· ····················
Q 5897 hCoV-19/England/MILK-A6A732/2020|EPI_ISL_627516|20 ···················· ···················· ····················· ···················· ······T············· ····················
Q 5898 hCoV-19/England/MILK-A6A78D/2020|EPI_ISL_627517|20 ···················· ···················· ····················· ···················· ···················· ····················
Q 5899 hCoV-19/Northern Ireland/NIRE-108993/2020|EPI_ISL_ ···················· ···················· ····················· ···················· ···················· ····················
Q 5900 hCoV-19/Northern Ireland/NIRE-1089A2/2020|EPI_ISL_ ···················· ···················· ····················· ···················· ···················· ····················
Q 5901 hCoV-19/Northern Ireland/NIRE-1089C0/2020|EPI_ISL_ ···················· ···················· ····················· ···················· ···················· ····················
Q 5902 hCoV-19/Northern Ireland/NIRE-1089DF/2020|EPI_ISL_ ···················· ···················· ····················· ···················· ···················· ····················
Q 5903 hCoV-19/Northern Ireland/NIRE-1089EE/2020|EPI_ISL_ ···················· ···················· ····················· ···················· ···················· ····················
Q 5904 hCoV-19/Northern Ireland/NIRE-1089FD/2020|EPI_ISL_ ···················· ···················· ····················· ···················· ···················· ····················
Q 5905 hCoV-19/Northern Ireland/NIRE-108A09/2020|EPI_ISL_ ···················· ···················· ····················· ···················· ···················· ····················
Q 5906 hCoV-19/Northern Ireland/NIRE-108A18/2020|EPI_ISL_ ···················· ···················· ····················· ···················· ···················· ····················
Q 5907 hCoV-19/Northern Ireland/NIRE-108A27/2020|EPI_ISL_ ···················· ···················· ····················· ···················· ···················· ····················
Q 5908 hCoV-19/Northern Ireland/NIRE-108A45/2020|EPI_ISL_ ···················· ···················· ····················· ···················· ···················· ····················
Q 5909 hCoV-19/Northern Ireland/NIRE-108A54/2020|EPI_ISL_ ···················· ···················· ····················· ···················· ···················· ····················
Q 5910 hCoV-19/Northern Ireland/NIRE-108A63/2020|EPI_ISL_ ···················· ···················· ····················· ···················· ···················· ····················
Q 5911 hCoV-19/USA/MI-UM-10036488294/2020|EPI_ISL_628839| ···················· ···················· ····················· ···················· ···················· ····················
Q 5912 hCoV-19/USA/MI-UM-10036464027/2020|EPI_ISL_628840| ···················· ···················· ····················· ···················· ···················· ····················
Q 5913 hCoV-19/USA/MI-UM-10036485874/2020|EPI_ISL_628841| ···················· ···················· ····················· ···················· ···················· ····················
Q 5914 hCoV-19/USA/MI-UM-10036496609/2020|EPI_ISL_628842| ···················· ·············A······ ····················· ···················· ···················· ····················
Q 5915 hCoV-19/USA/MI-UM-10036455460/2020|EPI_ISL_628843| ···················· ···················· ····················· ···················· ···················· ····················
Q 5916 hCoV-19/USA/MI-UM-10036496672/2020|EPI_ISL_628844| ···················· ···················· ····················· ···················· ···················· ····················
Q 5917 hCoV-19/USA/MI-UM-10036470386/2020|EPI_ISL_628845| ···················· ···················· ····················· ···················· ···················· ····················
Q 5918 hCoV-19/USA/MI-UM-10036455304/2020|EPI_ISL_628846| ···················· ···················· ····················· ···················· ···················· ····················
Q 5921 hCoV-19/USA/MI-UM-10036483226/2020|EPI_ISL_628849| ···················· ·A·················· ····················· ···················· ···················· ····················
Q 5922 hCoV-19/USA/MI-UM-10036466389/2020|EPI_ISL_628851| ···················· ···················· ····················· ···················· ···················· ····················
Q 5923 hCoV-19/USA/MI-UM-10036456115/2020|EPI_ISL_628853| ···················· ···················· ····················· ···················· ···················· ····················
Q 5925 hCoV-19/USA/MI-UM-10036495313/2020|EPI_ISL_628857| ···················· ···················· ····················· ···················· ···················· ····················
Q 5926 hCoV-19/USA/MI-UM-10036496799/2020|EPI_ISL_628859| ···················· ·A·················· ····················· ···················· ···················· ····················
Q 5928 hCoV-19/USA/NY-NYCPHL-000683/2020|EPI_ISL_632100|2 ···················· ···················· ····················· ···················· ···················· ····················
Q 5929 hCoV-19/USA/NY-NYCPHL-000684/2020|EPI_ISL_632101|2 ···················· ···················· ····················· ···················· ···················· ····················
Q 5930 hCoV-19/USA/NY-NYCPHL-000685/2020|EPI_ISL_632102|2 ···················· ···················· ····················· ···················· ···················· ····················
Q 5931 hCoV-19/USA/NY-NYCPHL-000686/2020|EPI_ISL_632103|2 ···················· ···················· ····················· ···················· ···················· ····················
Q 5932 hCoV-19/USA/NY-NYCPHL-000687/2020|EPI_ISL_632104|2 ···················· ···················· ····················· ···················· ···················· ····················

| | | | | | | | |
|---|---|---|---|---|---|---|---|
| Q | 5933 hCoV-19/USA/NY-NYCPHL-000689/2020\|EPI_ISL_632105\|2 | ···················· | ···················· | ····················· | ···················· | ···················· | ···················· |
| Q | 5934 hCoV-19/USA/NY-NYCPHL-000690/2020\|EPI_ISL_632106\|2 | ···················· | ···················· | ····················· | ···················· | ···················· | ···················· |
| Q | 5935 hCoV-19/USA/NY-NYCPHL-000691/2020\|EPI_ISL_632107\|2 | ···················· | ···················· | ····················· | ···················· | ···················· | ···················· |
| Q | 5936 hCoV-19/USA/NY-NYCPHL-000692/2020\|EPI_ISL_632108\|2 | ···················· | ···················· | ····················· | ···················· | ···················· | ···················· |
| Q | 5937 hCoV-19/USA/NY-NYCPHL-000693/2020\|EPI_ISL_632109\|2 | ···················· | ···················· | ····················· | ···················· | ···················· | ···················· |
| Q | 5938 hCoV-19/USA/NY-NYCPHL-000694/2020\|EPI_ISL_632110\|2 | ···················· | ···················· | ····················· | ···················· | ···················· | ···················· |
| Q | 5939 hCoV-19/USA/NY-NYCPHL-000695/2020\|EPI_ISL_632111\|2 | ···················· | ···················· | ····················· | ···················· | ···················· | ···················· |
| Q | 5940 hCoV-19/USA/NY-NYCPHL-000730/2020\|EPI_ISL_632112\|2 | ···················· | ···················· | ····················· | ···················· | ···················· | ···················· |
| Q | 5941 hCoV-19/USA/NY-NYCPHL-000731/2020\|EPI_ISL_632113\|2 | ···················· | ···················· | ····················· | ···················· | ···················· | ···················· |
| Q | 5942 hCoV-19/USA/NY-NYCPHL-000732/2020\|EPI_ISL_632114\|2 | ···················· | ···················· | ····················· | ···················· | ···················· | ···················· |
| Q | 5943 hCoV-19/USA/NY-NYCPHL-000733/2020\|EPI_ISL_632115\|2 | ···················· | ···················· | ····················· | ···················· | ···················· | ···················· |
| Q | 5944 hCoV-19/USA/NY-NYCPHL-000734/2020\|EPI_ISL_632116\|2 | ···················· | ···················· | ····················· | ···················· | ···················· | ···················· |
| Q | 5945 hCoV-19/USA/NY-NYCPHL-000735/2020\|EPI_ISL_632117\|2 | ···················· | ···················· | ····················· | ···················· | ···················· | ···················· |
| Q | 5946 hCoV-19/USA/NY-NYCPHL-000737/2020\|EPI_ISL_632118\|2 | ···················· | ···················· | ····················· | ···················· | ···················· | ···················· |
| Q | 5947 hCoV-19/USA/NY-NYCPHL-000738/2020\|EPI_ISL_632119\|2 | ···················· | ···················· | ····················· | ···················· | ···················· | ···················· |
| Q | 5948 hCoV-19/USA/NY-NYCPHL-000739/2020\|EPI_ISL_632120\|2 | ···················· | ···················· | ····················· | ···················· | ···················· | ···················· |
| Q | 5949 hCoV-19/USA/NY-NYCPHL-000740/2020\|EPI_ISL_632121\|2 | ···················· | ···················· | ····················· | ···················· | ···················· | ···················· |
| Q | 5950 hCoV-19/USA/NY-NYCPHL-000741/2020\|EPI_ISL_632122\|2 | ···················· | ···················· | ····················· | ···················· | ···················· | ···················· |
| Q | 5951 hCoV-19/USA/NY-NYCPHL-000742/2020\|EPI_ISL_632123\|2 | ···················· | ···················· | ····················· | ···················· | ···················· | ···················· |
| Q | 5952 hCoV-19/USA/NY-NYCPHL-000743/2020\|EPI_ISL_632124\|2 | ···················· | ···················· | ····················· | ···················· | ···················· | ···················· |
| Q | 5953 hCoV-19/USA/NY-NYCPHL-000744/2020\|EPI_ISL_632125\|2 | ···················· | ···················· | ····················· | ···················· | ···················· | ···················· |
| Q | 5954 hCoV-19/USA/NY-NYCPHL-000745/2020\|EPI_ISL_632126\|2 | ···················· | ···················· | ····················· | ···················· | ···················· | ···················· |
| Q | 5955 hCoV-19/USA/NY-NYCPHL-000746/2020\|EPI_ISL_632127\|2 | ···················· | ···················· | ····················· | ···················· | ···················· | ···················· |
| Q | 5956 hCoV-19/USA/NY-NYCPHL-000748/2020\|EPI_ISL_632128\|2 | ···················· | ···················· | ····················· | ···················· | ···················· | ···················· |
| Q | 5957 hCoV-19/USA/NY-NYCPHL-000749/2020\|EPI_ISL_632129\|2 | ···················· | ···················· | ····················· | ···················· | ···················· | ···················· |
| Q | 5958 hCoV-19/USA/NY-NYCPHL-000750/2020\|EPI_ISL_632130\|2 | ···················· | ···················· | ····················· | ···················· | ···················· | ···················· |
| Q | 5959 hCoV-19/USA/NY-NYCPHL-000751/2020\|EPI_ISL_632131\|2 | ···················· | ···················· | ····················· | ···················· | ···················· | ···················· |
| Q | 5960 hCoV-19/USA/NY-NYCPHL-000752/2020\|EPI_ISL_632132\|2 | ···················· | ···················· | ····················· | ···················· | ···················· | ···················· |
| Q | 5961 hCoV-19/USA/NY-NYCPHL-000753/2020\|EPI_ISL_632133\|2 | ···················· | ···················· | ····················· | ···················· | ···················· | ···················· |
| Q | 5962 hCoV-19/USA/NY-NYCPHL-000754/2020\|EPI_ISL_632134\|2 | ···················· | ···················· | ····················· | ···················· | ···················· | ···················· |
| Q | 5963 hCoV-19/USA/NY-NYCPHL-000755/2020\|EPI_ISL_632135\|2 | ···················· | ···················· | ····················· | ···················· | ···················· | ···················· |
| Q | 5964 hCoV-19/USA/NY-NYCPHL-000756/2020\|EPI_ISL_632136\|2 | ···················· | ···················· | ····················· | ···················· | ···················· | ···················· |
| Q | 5965 hCoV-19/USA/NY-NYCPHL-000757/2020\|EPI_ISL_632137\|2 | ···················· | ···················· | ····················· | ···················· | ···················· | ···················· |
| Q | 5966 hCoV-19/USA/NY-NYCPHL-000758/2020\|EPI_ISL_632138\|2 | ···················· | ···················· | ····················· | ···················· | ···················· | ···················· |
| Q | 5967 hCoV-19/USA/NY-NYCPHL-000759/2020\|EPI_ISL_632139\|2 | ···················· | ···················· | ····················· | ···················· | ···················· | ···················· |
| Q | 5968 hCoV-19/USA/NY-NYCPHL-000760/2020\|EPI_ISL_632140\|2 | ···················· | ···················· | ····················· | ···················· | ···················· | ···················· |
| Q | 5969 hCoV-19/USA/NY-NYCPHL-000761/2020\|EPI_ISL_632141\|2 | ···················· | ···················· | ····················· | ···················· | ···················· | ···················· |
| Q | 5970 hCoV-19/USA/NY-NYCPHL-000762/2020\|EPI_ISL_632142\|2 | ···················· | ···················· | ····················· | ···················· | ···················· | ···················· |
| Q | 5971 hCoV-19/USA/NY-NYCPHL-000763/2020\|EPI_ISL_632143\|2 | ···················· | ···················· | ····················· | ···················· | ···················· | ···················· |
| Q | 5972 hCoV-19/USA/NY-NYCPHL-000764/2020\|EPI_ISL_632144\|2 | ···················· | ···················· | ····················· | ···················· | ···················· | ···················· |
| Q | 5973 hCoV-19/USA/NY-NYCPHL-000765/2020\|EPI_ISL_632145\|2 | ···················· | ···················· | ····················· | ···················· | ···················· | ···················· |
| Q | 5974 hCoV-19/USA/NY-NYCPHL-000766/2020\|EPI_ISL_632146\|2 | ···················· | ··············C····· | ····················· | ···················· | ···················· | ···················· |
| Q | 5975 hCoV-19/USA/NY-NYCPHL-000767/2020\|EPI_ISL_632147\|2 | ···················· | ···················· | ····················· | ···················· | ···················· | ···················· |
| Q | 5976 hCoV-19/USA/NY-NYCPHL-000768/2020\|EPI_ISL_632148\|2 | ···················· | ···················· | ····················· | ···················· | ···················· | ···················· |
| Q | 5977 hCoV-19/USA/NY-NYCPHL-000769/2020\|EPI_ISL_632149\|2 | ···················· | ···················· | ····················· | ···················· | ···················· | ···················· |
| Q | 5978 hCoV-19/USA/NY-NYCPHL-000770/2020\|EPI_ISL_632150\|2 | ···················· | ···················· | ····················· | ···················· | ···················· | ···················· |
| Q | 5979 hCoV-19/USA/NY-NYCPHL-000771/2020\|EPI_ISL_632151\|2 | ···················· | ···················· | ····················· | ···················· | ···················· | ···················· |
| Q | 5980 hCoV-19/USA/NY-NYCPHL-000772/2020\|EPI_ISL_632152\|2 | ···················· | ···················· | ····················· | ···················· | ···················· | ···················· |
| Q | 5981 hCoV-19/USA/NY-NYCPHL-000773/2020\|EPI_ISL_632153\|2 | ···················· | ···················· | ····················· | ···················· | ···················· | ···················· |
| Q | 5982 hCoV-19/USA/NY-NYCPHL-000774/2020\|EPI_ISL_632154\|2 | ···················· | ···················· | ····················· | ···················· | ···················· | ···················· |
| Q | 5983 hCoV-19/USA/NY-NYCPHL-000775/2020\|EPI_ISL_632155\|2 | ···················· | ···················· | ····················· | ···················· | ···················· | ···················· |
| Q | 5984 hCoV-19/USA/NY-NYCPHL-000776/2020\|EPI_ISL_632156\|2 | ···················· | ···················· | ····················· | ···················· | ···················· | ···················· |
| Q | 5985 hCoV-19/USA/NY-NYCPHL-000777/2020\|EPI_ISL_632157\|2 | ···················· | ···················· | ····················· | ···················· | ···················· | ···················· |
| Q | 5986 hCoV-19/USA/NY-NYCPHL-000778/2020\|EPI_ISL_632158\|2 | ···················· | ···················· | ····················· | ···················· | ···················· | ···················· |
| Q | 5987 hCoV-19/USA/NY-NYCPHL-000779/2020\|EPI_ISL_632159\|2 | ···················· | ···················· | ····················· | ···················· | ···················· | ···················· |
| Q | 5988 hCoV-19/USA/NY-NYCPHL-000780/2020\|EPI_ISL_632160\|2 | ···················· | ···················· | ····················· | ···················· | ···················· | ···················· |
| Q | 5989 hCoV-19/USA/NY-NYCPHL-000781/2020\|EPI_ISL_632161\|2 | ···················· | ···················· | ····················· | ···················· | ···················· | ···················· |
| Q | 5990 hCoV-19/USA/NY-NYCPHL-000782/2020\|EPI_ISL_632162\|2 | ···················· | ···················· | ····················· | ···················· | ···················· | ···················· |
| Q | 5991 hCoV-19/USA/NY-NYCPHL-000783/2020\|EPI_ISL_632163\|2 | ···················· | ···················· | ····················· | ···················· | ···················· | ···················· |
| Q | 5992 hCoV-19/USA/NY-NYCPHL-000784/2020\|EPI_ISL_632164\|2 | ···················· | ···················· | ····················· | ···················· | ···················· | ···················· |
| Q | 5993 hCoV-19/USA/NY-NYCPHL-000785/2020\|EPI_ISL_632165\|2 | ···················· | ···················· | ····················· | ···················· | ···················· | ···················· |
| Q | 5994 hCoV-19/USA/NY-NYCPHL-000786/2020\|EPI_ISL_632166\|2 | ···················· | ···················· | ····················· | ···················· | ···················· | ···················· |
| Q | 5995 hCoV-19/USA/NY-NYCPHL-000787/2020\|EPI_ISL_632167\|2 | ···················· | ···················· | ····················· | ···················· | ···················· | ···················· |
| Q | 5996 hCoV-19/USA/NY-NYCPHL-000788/2020\|EPI_ISL_632168\|2 | ···················· | ···················· | ····················· | ···················· | ···················· | ···················· |
| Q | 5997 hCoV-19/USA/NY-NYCPHL-000789/2020\|EPI_ISL_632169\|2 | ···················· | ···················· | ····················· | ···················· | ···················· | ···················· |
| Q | 5998 hCoV-19/USA/NY-NYCPHL-000790/2020\|EPI_ISL_632170\|2 | ···················· | ···················· | ····················· | ···················· | ···················· | ···················· |
| Q | 5999 hCoV-19/USA/NY-NYCPHL-000791/2020\|EPI_ISL_632171\|2 | ···················· | ···················· | ····················· | ···················· | ···················· | ···················· |
| Q | 6000 hCoV-19/USA/NY-NYCPHL-000792/2020\|EPI_ISL_632172\|2 | ···················· | ···················· | ····················· | ···················· | ···················· | ···················· |
| Q | 6001 hCoV-19/USA/NY-NYCPHL-000793/2020\|EPI_ISL_632173\|2 | ···················· | ···················· | ····················· | ···················· | ···················· | ···················· |
| Q | 6002 hCoV-19/USA/NY-NYCPHL-000794/2020\|EPI_ISL_632174\|2 | ···················· | ···················· | ····················· | ···················· | ···················· | ···················· |
| Q | 6003 hCoV-19/USA/NY-NYCPHL-000795/2020\|EPI_ISL_632175\|2 | ···················· | ···················· | ····················· | ···················· | ···················· | ···················· |
| Q | 6004 hCoV-19/USA/NY-NYCPHL-000796/2020\|EPI_ISL_632176\|2 | ···················· | ···················· | ····················· | ···················· | ···················· | ···················· |
| Q | 6005 hCoV-19/USA/NY-NYCPHL-000797/2020\|EPI_ISL_632177\|2 | ···················· | ···················· | ····················· | ···················· | ···················· | ···················· |
| Q | 6006 hCoV-19/USA/NY-NYCPHL-000798/2020\|EPI_ISL_632178\|2 | ···················· | ···················· | ····················· | ···················· | ···················· | ···················· |
| Q | 6007 hCoV-19/USA/NY-NYCPHL-000799/2020\|EPI_ISL_632179\|2 | ···················· | ···················· | ····················· | ···················· | ···················· | ···················· |
| Q | 6008 hCoV-19/England/QEUH-AE4FED/2020\|EPI_ISL_633404\|20 | ···················· | ···················· | ····················· | ···················· | ···················· | ···················· |
| Q | 6009 hCoV-19/England/CAMC-AEA8FB/2020\|EPI_ISL_633405\|20 | ···················· | ···················· | ····················· | ···················· | ······T············· | ···················· |
| Q | 6010 hCoV-19/England/QEUH-AE5298/2020\|EPI_ISL_633406\|20 | ···················· | ···················· | ····················· | ···················· | ···················· | ···A················ |
| Q | 6011 hCoV-19/England/CAMC-AEAC68/2020\|EPI_ISL_633407\|20 | ···················· | ···················· | ····················· | ···················· | ···················· | ···················· |
| Q | 6012 hCoV-19/England/CAMC-AEAC4A/2020\|EPI_ISL_633408\|20 | ···················· | ···················· | ····················· | ···················· | ···················· | ···················· |

```
Q 6013 hCoV-19/England/QEUH-AE237A/2020|EPI_ISL_633409|20 ···················· ···················· ···················· ···················· ···················· ····················
Q 6014 hCoV-19/England/QEUH-AE2723/2020|EPI_ISL_633410|20 ···················· ···················· ···················· ···················· ···················· ····················
Q 6015 hCoV-19/England/QEUH-AE30FA/2020|EPI_ISL_633411|20 ···················· ···················· ···················· ···················· ···················· ····················
Q 6016 hCoV-19/England/QEUH-AE1B64/2020|EPI_ISL_633412|20 ···················· ···················· ···················· ···················· ···················· ····················
Q 6017 hCoV-19/England/QEUH-AE1F77/2020|EPI_ISL_633413|20 ···················· ···················· ···················· ···················· ···················· ····················
Q 6018 hCoV-19/England/QEUH-AE1C9E/2020|EPI_ISL_633414|20 ···················· ···················· ···················· ···················· ···················· ····················
Q 6019 hCoV-19/England/QEUH-AE28E4/2020|EPI_ISL_633416|20 ···················· ···················· ···················· ···················· ···················· ····················
Q 6020 hCoV-19/England/QEUH-AE27AB/2020|EPI_ISL_633426|20 ···················· ···················· ···················· ···················· ···················· ····················
Q 6021 hCoV-19/England/QEUH-AE232F/2020|EPI_ISL_633427|20 ···················· ···················· ···················· ···················· ···················· ····················
Q 6022 hCoV-19/England/QEUH-AE2C51/2020|EPI_ISL_633428|20 ···················· ···················· ···················· ···················· ···················· ····················
Q 6023 hCoV-19/England/QEUH-AE2680/2020|EPI_ISL_633429|20 ···················· ···················· ···················· ···················· ···················· ····················
Q 6024 hCoV-19/England/QEUH-AE2C15/2020|EPI_ISL_633430|20 ···················· ···················· ···················· ···················· ···················· ····················
Q 6025 hCoV-19/England/QEUH-AE2D30/2020|EPI_ISL_633431|20 ···················· ···················· ···················· ···················· ···················· ····················
Q 6026 hCoV-19/England/QEUH-AE2741/2020|EPI_ISL_633433|20 ···················· ···················· ···················· ···················· ···················· ····················
Q 6027 hCoV-19/England/QEUH-AE29C3/2020|EPI_ISL_633434|20 ···················· ···················· ···················· ···················· ···················· ····················
Q 6028 hCoV-19/England/QEUH-AE235C/2020|EPI_ISL_633435|20 ···················· ···················· ···················· ···················· ···················· ····················
Q 6029 hCoV-19/England/QEUH-AE1B73/2020|EPI_ISL_633436|20 ···················· ···················· ···················· ···················· ···················· ····················
Q 6030 hCoV-19/England/QEUH-AE1C16/2020|EPI_ISL_633437|20 ···················· ···················· ···················· ···················· ···················· ····················
Q 6031 hCoV-19/England/QEUH-AE27F6/2020|EPI_ISL_633438|20 ···················· ···················· ···················· ···················· ···················· ····················
Q 6032 hCoV-19/England/QEUH-AE2037/2020|EPI_ISL_633439|20 ···················· ···················· ···················· ···················· ···················· ····················
Q 6033 hCoV-19/England/QEUH-AE26BD/2020|EPI_ISL_633440|20 ···················· ···················· ···················· ···················· ···················· ····················
Q 6034 hCoV-19/England/QEUH-AE28A8/2020|EPI_ISL_633441|20 ···················· ···················· ···················· ···················· ···················· ····················
Q 6035 hCoV-19/England/QEUH-AE2B54/2020|EPI_ISL_633442|20 ···················· ·A·················· ···················· ···················· ···················· ····················
Q 6036 hCoV-19/England/QEUH-AE2D03/2020|EPI_ISL_633444|20 ···················· ···················· ···················· ···················· ···················· ····················
Q 6037 hCoV-19/England/QEUH-AE2A84/2020|EPI_ISL_633445|20 ···················· ·A·················· ···················· ···················· ···················· ··A·················
Q 6038 hCoV-19/England/QEUH-AE285D/2020|EPI_ISL_633446|20 ···················· ···················· ···················· ···················· ···················· ····················
Q 6039 hCoV-19/England/QEUH-AE2EB5/2020|EPI_ISL_633447|20 ···················· ···················· ···················· ···················· ···················· ····················
Q 6040 hCoV-19/England/QEUH-AE2E5B/2020|EPI_ISL_633448|20 ···················· ···················· ···················· ···················· ···················· ····················
Q 6041 hCoV-19/England/QEUH-AE283F/2020|EPI_ISL_633449|20 ···················· ···················· ···················· ···················· ···················· ····················
Q 6042 hCoV-19/England/QEUH-AE1C70/2020|EPI_ISL_633450|20 ···················· ···················· ···················· ···················· ···················· ····················
Q 6043 hCoV-19/England/QEUH-AE2D8B/2020|EPI_ISL_633451|20 ···················· ···················· ···················· ···················· ···················· ····················
Q 6044 hCoV-19/England/QEUH-AE2BAF/2020|EPI_ISL_633452|20 ···················· ···················· ···················· ···················· ···················· ····················
Q 6045 hCoV-19/England/QEUH-AE2310/2020|EPI_ISL_633453|20 ···················· ···················· ···················· ···················· ···················· ····················
Q 6046 hCoV-19/England/QEUH-AE269F/2020|EPI_ISL_633454|20 ···················· ···················· ···················· ···················· ···················· ····················
Q 6047 hCoV-19/England/QEUH-AE2A75/2020|EPI_ISL_633455|20 ···················· ···················· ···················· ···················· ···················· ····················
Q 6048 hCoV-19/England/QEUH-AE2107/2020|EPI_ISL_633456|20 ···················· ·A·················· ···················· ···················· ···················· ··A·················
Q 6049 hCoV-19/England/QEUH-AE242C/2020|EPI_ISL_633457|20 ···················· ···················· ···················· ···················· ···················· ····················
Q 6050 hCoV-19/England/QEUH-AE2BDC/2020|EPI_ISL_633458|20 ···················· ···················· ···················· ···················· ···················· ····················
Q 6051 hCoV-19/England/QEUH-AE27C9/2020|EPI_ISL_633459|20 ···················· ···················· ···················· ···················· ···················· ····················
Q 6052 hCoV-19/England/QEUH-AE2A48/2020|EPI_ISL_633460|20 ···················· ···················· ···················· ···················· ···················· ····················
Q 6053 hCoV-19/England/QEUH-AE2D5E/2020|EPI_ISL_633461|20 ···················· ···················· ···················· ···················· ···················· ····················
Q 6054 hCoV-19/England/QEUH-AE2DD6/2020|EPI_ISL_633462|20 ···················· ···················· ···················· ···················· ···················· ····················
Q 6055 hCoV-19/England/QEUH-AE21DA/2020|EPI_ISL_633464|20 ···················· ···················· ···················· ···················· ···················· ····················
Q 6056 hCoV-19/England/QEUH-AE2A39/2020|EPI_ISL_633466|20 ···················· ···················· ···················· ···················· ···················· ····················
Q 6057 hCoV-19/England/QEUH-AE26EA/2020|EPI_ISL_633467|20 ···················· ···················· ···················· ···················· ···················· ····················
Q 6058 hCoV-19/England/QEUH-AE2152/2020|EPI_ISL_633468|20 ···················· ···················· ···················· ···················· ···················· ····················
Q 6059 hCoV-19/England/QEUH-AE3081/2020|EPI_ISL_633469|20 ···················· ···················· ···················· ···················· ···················· ····················
Q 6060 hCoV-19/England/QEUH-AE2BBE/2020|EPI_ISL_633470|20 ···················· ···················· ···················· ···················· ···················· ····················
Q 6061 hCoV-19/England/QEUH-AE1E7A/2020|EPI_ISL_633471|20 ···················· ···················· ···················· ···················· ···················· ····················
Q 6062 hCoV-19/England/QEUH-AE1DD7/2020|EPI_ISL_633472|20 ···················· ···················· ···················· ···················· ···················· ····················
Q 6063 hCoV-19/England/QEUH-AE28C6/2020|EPI_ISL_633473|20 ···················· ···················· ···················· ···················· ···················· ····················
Q 6064 hCoV-19/England/QEUH-AE26CC/2020|EPI_ISL_633475|20 ···················· ···················· ···················· ···················· ···················· ····················
Q 6065 hCoV-19/England/QEUH-AE243B/2020|EPI_ISL_633476|20 ···················· ···················· ···················· ···················· ···················· ····················
Q 6066 hCoV-19/England/QEUH-AE1D9B/2020|EPI_ISL_633477|20 ···················· ···················· ···················· ···················· ···················· ····················
Q 6067 hCoV-19/England/QEUH-AE2802/2020|EPI_ISL_633478|20 ···················· ···················· ···················· ···················· ···················· ··A·················
Q 6068 hCoV-19/England/QEUH-AE24A4/2020|EPI_ISL_633479|20 ···················· ···················· ···················· ···················· ···················· ····················
Q 6069 hCoV-19/England/QEUH-AE1D8C/2020|EPI_ISL_633480|20 ···················· ···················· ···················· ···················· ···················· ····················
Q 6070 hCoV-19/England/QEUH-AE2028/2020|EPI_ISL_633481|20 ···················· ···················· ···················· ···················· ···················· ····················
Q 6071 hCoV-19/England/QEUH-AE236B/2020|EPI_ISL_633482|20 ···················· ···················· ···················· ···················· ···················· ····················
Q 6072 hCoV-19/England/QEUH-AE2CE8/2020|EPI_ISL_633483|20 ···················· ···················· ···················· ···················· ···················· ····················
Q 6073 hCoV-19/England/QEUH-AE2583/2020|EPI_ISL_633484|20 ···················· ···················· ···················· ···················· ···················· ····················
Q 6074 hCoV-19/England/QEUH-AE2046/2020|EPI_ISL_633485|20 ···················· ···················· ···················· ···················· ···················· ····················
Q 6075 hCoV-19/England/QEUH-AE2ED3/2020|EPI_ISL_633486|20 ···················· ···················· ···················· ···················· ···················· ····················
Q 6076 hCoV-19/England/QEUH-AE2653/2020|EPI_ISL_633487|20 ···················· ···················· ···················· ···················· ···················· ····················
Q 6077 hCoV-19/England/QEUH-AE23F2/2020|EPI_ISL_633488|20 ···················· ···················· ···················· ···················· ···················· ····················
Q 6078 hCoV-19/England/QEUH-AE2F3A/2020|EPI_ISL_633490|20 ···················· ···················· ···················· ···················· ···················· ····················
Q 6079 hCoV-19/England/QEUH-AE22B9/2020|EPI_ISL_633491|20 ···················· ···················· ···················· ···················· ···················· ····················
Q 6080 hCoV-19/England/QEUH-AE26F9/2020|EPI_ISL_633493|20 ···················· ···················· ···················· ···················· ···················· ····················
Q 6081 hCoV-19/England/QEUH-AE2D7C/2020|EPI_ISL_633494|20 ···················· ···················· ···················· ···················· ···················· ····················
Q 6082 hCoV-19/England/QEUH-AE22AA/2020|EPI_ISL_633495|20 ···················· ···················· ···················· ···················· ···················· ····················
Q 6083 hCoV-19/England/QEUH-AE3063/2020|EPI_ISL_633496|20 ···················· ···················· ···················· ···················· ···················· ····················
Q 6084 hCoV-19/England/QEUH-AE2ADF/2020|EPI_ISL_633497|20 ···················· ···················· ···················· ···················· ···················· ····················
Q 6085 hCoV-19/England/QEUH-AE1FFF/2020|EPI_ISL_633499|20 ···················· ···················· ···················· ···················· ···················· ····················
Q 6086 hCoV-19/England/QEUH-AD43A8/2020|EPI_ISL_634700|20 ···················· ·A·················· ···················· ···················· ···················· ··A·················
Q 6087 hCoV-19/England/QEUH-AD1EC8/2020|EPI_ISL_634701|20 ···················· ···················· ···················· ···················· ···················· ····················
Q 6088 hCoV-19/England/QEUH-AD4478/2020|EPI_ISL_634702|20 ···················· ···················· ···················· ···················· ···················· ····················
Q 6089 hCoV-19/England/QEUH-AD1F10/2020|EPI_ISL_634703|20 ···················· ···················· ···················· ···················· ···················· ····················
Q 6090 hCoV-19/England/QEUH-AD46EB/2020|EPI_ISL_634705|20 ···················· ···················· ···················· ···················· ···················· ····················
Q 6091 hCoV-19/England/QEUH-AD5A0C/2020|EPI_ISL_634706|20 ···················· ·A·················· ···················· ···················· ···················· ····················
Q 6092 hCoV-19/England/QEUH-AD5C7F/2020|EPI_ISL_634708|20 ···················· ···················· ···················· ···················· ···················· ····················
```

| | | | | | | | | |
|---|---|---|---|---|---|---|---|---|
| Q | 6093 hCoV-19/England/QEUH-AD585D/2020\|EPI_ISL_634709\|20 | ···················· | ···················· | ···················· | ···················· | ···················· | ···················· | |
| Q | 6094 hCoV-19/England/QEUH-AD1FA7/2020\|EPI_ISL_634710\|20 | ···················· | ···················· | ···················· | ···················· | ···················· | ···················· | |
| Q | 6095 hCoV-19/England/QEUH-AD1639/2020\|EPI_ISL_634711\|20 | ···················· | ···················· | ···················· | ···················· | ···················· | ···················· | |
| Q | 6096 hCoV-19/England/QEUH-AD3EE4/2020\|EPI_ISL_634712\|20 | ···················· | ···················· | ···················· | ···················· | ···················· | ···················· | |
| Q | 6097 hCoV-19/England/QEUH-AD1A97/2020\|EPI_ISL_634735\|20 | ···················· | ···················· | ···················· | ···················· | ···················· | ···················· | |
| Q | 6098 hCoV-19/Wales/QEUH-AD17EB/2020\|EPI_ISL_634736\|2020 | ···················· | ·A·················· | ···················· | ···················· | ···················· | ···················· | |
| Q | 6099 hCoV-19/Wales/QEUH-AD3DC9/2020\|EPI_ISL_634738\|2020 | ···················· | ···················· | ···················· | ···················· | ···················· | ···················· | |
| Q | 6100 hCoV-19/Wales/QEUH-AD251D/2020\|EPI_ISL_634739\|2020 | ···················· | ···················· | ···················· | ···················· | ···················· | ···················· | |
| Q | 6101 hCoV-19/Wales/QEUH-AD3D41/2020\|EPI_ISL_634740\|2020 | ···················· | ···················· | ···················· | ···················· | ···················· | ···················· | |
| Q | 6102 hCoV-19/Wales/QEUH-AD3A3B/2020\|EPI_ISL_634741\|2020 | ···················· | ···················· | ···················· | ···················· | ···················· | ···················· | |
| Q | 6103 hCoV-19/Wales/QEUH-AD33C7/2020\|EPI_ISL_634742\|2020 | ···················· | ···················· | ···················· | ···················· | ···················· | ···················· | |
| Q | 6104 hCoV-19/Wales/QEUH-AD1860/2020\|EPI_ISL_634744\|2020 | ···················· | ···················· | ···················· | ···················· | ···················· | ···················· | |
| Q | 6105 hCoV-19/Wales/QEUH-AD1790/2020\|EPI_ISL_634745\|2020 | ···················· | ···················· | ···················· | ···················· | ···················· | ···················· | |
| Q | 6106 hCoV-19/Wales/QEUH-AD3E5D/2020\|EPI_ISL_634746\|2020 | ···················· | ···················· | ···················· | ···················· | ···················· | ···················· | |
| Q | 6107 hCoV-19/Wales/QEUH-AD3B0B/2020\|EPI_ISL_634747\|2020 | ···················· | ···················· | ···················· | ···················· | ···················· | ···················· | |
| Q | 6108 hCoV-19/Wales/ALDP-AA3023/2020\|EPI_ISL_634758\|2020 | ···················· | ···················· | ···················· | ···················· | ···················· | ···················· | |
| Q | 6109 hCoV-19/Wales/ALDP-AA1C30/2020\|EPI_ISL_634759\|2020 | ···················· | ···················· | ···················· | ···················· | ···················· | ···················· | |
| Q | 6110 hCoV-19/Wales/ALDP-AA24DD/2020\|EPI_ISL_634760\|2020 | ···················· | ···················· | ···················· | ···················· | ···················· | ···················· | |
| Q | 6111 hCoV-19/Wales/ALDP-AA308D/2020\|EPI_ISL_634761\|2020 | ···················· | ···················· | ···················· | ···················· | ···················· | ···················· | |
| Q | 6112 hCoV-19/Wales/ALDP-AA2525/2020\|EPI_ISL_634762\|2020 | ···················· | ···················· | ···················· | ···················· | ···················· | ···················· | |
| Q | 6113 hCoV-19/Wales/ALDP-AA1BE8/2020\|EPI_ISL_634763\|2020 | ···················· | ···················· | ···················· | ···················· | ···················· | ···················· | |
| Q | 6114 hCoV-19/Wales/ALDP-AA1C6D/2020\|EPI_ISL_634764\|2020 | ···················· | ···················· | ···················· | ···················· | ···················· | ···················· | |
| Q | 6115 hCoV-19/Wales/ALDP-AA25AD/2020\|EPI_ISL_634766\|2020 | ···················· | ···················· | ···················· | ···················· | ···················· | ···················· | |
| Q | 6116 hCoV-19/Wales/ALDP-AA3050/2020\|EPI_ISL_634767\|2020 | ···················· | ···················· | ···················· | ···················· | ···················· | ···················· | |
| Q | 6117 hCoV-19/Wales/ALDP-AA1B42/2020\|EPI_ISL_634768\|2020 | ···················· | ···················· | ···················· | ···················· | ···················· | ···················· | |
| Q | 6118 hCoV-19/Wales/ALDP-AA2570/2020\|EPI_ISL_634769\|2020 | ···················· | ···················· | ···················· | ···················· | ···················· | ···················· | |
| Q | 6119 hCoV-19/Wales/ALDP-AA258F/2020\|EPI_ISL_634770\|2020 | ···················· | ···················· | ···················· | ···················· | ···················· | ···················· | |
| Q | 6120 hCoV-19/Wales/ALDP-AA2543/2020\|EPI_ISL_634771\|2020 | ···················· | ···················· | ···················· | ···················· | ···················· | ···················· | |
| Q | 6121 hCoV-19/Wales/ALDP-AA2FAF/2020\|EPI_ISL_634772\|2020 | ···················· | ···················· | ···················· | ···················· | ···················· | ···················· | |
| Q | 6122 hCoV-19/Wales/ALDP-AA1C12/2020\|EPI_ISL_634773\|2020 | ···················· | ···················· | ···················· | ···················· | ···················· | ···················· | |
| Q | 6123 hCoV-19/Wales/ALDP-AA50B8/2020\|EPI_ISL_634774\|2020 | ···················· | ···················· | ···················· | ···················· | ···················· | ···················· | |
| Q | 6124 hCoV-19/Wales/ALDP-AA2534/2020\|EPI_ISL_634775\|2020 | ···················· | ···················· | ···················· | ···················· | ···················· | ···················· | |
| Q | 6125 hCoV-19/Wales/ALDP-AA24FB/2020\|EPI_ISL_634776\|2020 | ···················· | ···················· | ···················· | ···················· | ···················· | ···················· | |
| Q | 6126 hCoV-19/Wales/ALDP-AA259E/2020\|EPI_ISL_634777\|2020 | ···················· | ···················· | ···················· | ···················· | ···················· | ···················· | |
| Q | 6127 hCoV-19/Wales/ALDP-AA2FEB/2020\|EPI_ISL_634778\|2020 | ···················· | ···················· | ···················· | ···················· | ···················· | ···················· | |
| Q | 6128 hCoV-19/Wales/ALDP-AA1BCA/2020\|EPI_ISL_634781\|2020 | ···················· | ···················· | ···················· | ···················· | ··A················· | ···················· | |
| Q | 6129 hCoV-19/Wales/ALDP-AA24A0/2020\|EPI_ISL_634782\|2020 | ···················· | ···················· | ···················· | ···················· | ···················· | ···················· | |
| Q | 6130 hCoV-19/Wales/ALDP-AA2FDC/2020\|EPI_ISL_634783\|2020 | ···················· | ···················· | ···················· | ···················· | ···················· | ···················· | |
| Q | 6131 hCoV-19/Wales/ALDP-AA30AB/2020\|EPI_ISL_634784\|2020 | ···················· | ···················· | ···················· | ···················· | ···················· | ···················· | |
| Q | 6132 hCoV-19/Wales/ALDP-AA24BF/2020\|EPI_ISL_634785\|2020 | ···················· | ···················· | ···················· | ···················· | ···················· | ···················· | |
| Q | 6133 hCoV-19/Wales/ALDP-AA504F/2020\|EPI_ISL_634786\|2020 | ···················· | ···················· | ···················· | ···················· | ···················· | ···················· | |
| Q | 6134 hCoV-19/Wales/ALDP-AA2516/2020\|EPI_ISL_634788\|2020 | ···················· | ···················· | ···················· | ···················· | ···················· | ···················· | |
| Q | 6135 hCoV-19/Wales/CAMC-AAF83E/2020\|EPI_ISL_634790\|2020 | ···················· | ···················· | ···················· | ···················· | ···················· | ···················· | |
| Q | 6136 hCoV-19/Wales/CAMC-AAF86B/2020\|EPI_ISL_634791\|2020 | ···················· | ···················· | ···················· | ···················· | ···················· | ···················· | |
| Q | 6137 hCoV-19/Wales/CAMC-AAF9B3/2020\|EPI_ISL_634792\|2020 | ···················· | ···················· | ···················· | ···················· | ···················· | ···················· | |
| Q | 6138 hCoV-19/Wales/CAMC-AAF9C2/2020\|EPI_ISL_634793\|2020 | ···················· | ···················· | ···················· | ···················· | ···················· | ···················· | |
| Q | 6139 hCoV-19/Wales/CAMC-AAF87A/2020\|EPI_ISL_634794\|2020 | ···················· | ···················· | ···················· | ···················· | ···················· | ···················· | |
| Q | 6140 hCoV-19/Wales/CAMC-AAF6DA/2020\|EPI_ISL_634795\|2020 | ···················· | ···················· | ···················· | ···················· | ···················· | ···················· | |
| Q | 6141 hCoV-19/Wales/CAMC-AAF85C/2020\|EPI_ISL_634796\|2020 | ···················· | ···················· | ···················· | ···················· | ···················· | ···················· | |
| Q | 6142 hCoV-19/Wales/CAMC-AAF82F/2020\|EPI_ISL_634797\|2020 | ···················· | ···················· | ···················· | ···················· | ···················· | ···················· | |
| Q | 6143 hCoV-19/Wales/CAMC-AAF5CE/2020\|EPI_ISL_634798\|2020 | ···················· | ···················· | ···················· | ···················· | ···················· | ···················· | |
| Q | 6144 hCoV-19/Wales/CAMC-AAF84D/2020\|EPI_ISL_634799\|2020 | ···················· | ···················· | ···················· | ···················· | ···················· | ···················· | |
| Q | 6145 hCoV-19/England/MILK-A6A36B/2020\|EPI_ISL_627500\|20 | ···················· | ···················· | ···················· | ···················· | ···················· | ···················· | |
| Q | 6146 hCoV-19/England/MILK-A6A389/2020\|EPI_ISL_627501\|20 | ···················· | ···················· | ···················· | ···················· | ···················· | ···················· | |
| Q | 6147 hCoV-19/England/MILK-A6A3D4/2020\|EPI_ISL_627502\|20 | ···················· | ···················· | ···················· | ···················· | ···················· | ···················· | |
| Q | 6148 hCoV-19/Scotland/MILK-A6A3E3/2020\|EPI_ISL_627503\|2 | ···················· | ···················· | ···················· | ···················· | ···················· | ···················· | |
| Q | 6149 hCoV-19/England/MILK-A6A40E/2020\|EPI_ISL_627504\|20 | ···················· | ···················· | ···················· | ···················· | ···················· | ···················· | |
| Q | 6150 hCoV-19/England/MILK-A6A459/2020\|EPI_ISL_627505\|20 | ···················· | ·A·················· | ···················· | ···················· | ···················· | ···················· | |
| Q | 6151 hCoV-19/England/MILK-A6A468/2020\|EPI_ISL_627506\|20 | ···················· | ···················· | ···················· | ···················· | ···················· | ···················· | |
| Q | 6152 hCoV-19/England/MILK-A6A486/2020\|EPI_ISL_627507\|20 | ···················· | ···················· | ···················· | ···················· | ···················· | ···················· | |
| Q | 6153 hCoV-19/Northern Ireland/NIRE-108A72/2020\|EPI_ISL_ | ···················· | ···················· | ···················· | ···················· | ···················· | ···················· | |
| Q | 6154 hCoV-19/Northern Ireland/NIRE-108A90/2020\|EPI_ISL_ | ···················· | ···················· | ···················· | ···················· | ···················· | ···················· | |
| Q | 6155 hCoV-19/Northern Ireland/NIRE-108AAF/2020\|EPI_ISL_ | ···················· | ···················· | ···················· | ···················· | ···················· | ···················· | |
| Q | 6156 hCoV-19/Northern Ireland/NIRE-108ADC/2020\|EPI_ISL_ | ···················· | ···················· | ···················· | ···················· | ···················· | ···················· | |
| Q | 6157 hCoV-19/Northern Ireland/NIRE-108AEB/2020\|EPI_ISL_ | ···················· | ·A·················· | ···················· | ···················· | ···················· | ···················· | |
| Q | 6158 hCoV-19/Northern Ireland/NIRE-108AFA/2020\|EPI_ISL_ | ···················· | ···················· | ···················· | ···················· | ···················· | ···················· | |
| Q | 6159 hCoV-19/Northern Ireland/NIRE-108B06/2020\|EPI_ISL_ | ···················· | ···················· | ···················· | ···················· | ···················· | ···················· | |
| Q | 6160 hCoV-19/Northern Ireland/NIRE-108B15/2020\|EPI_ISL_ | ···················· | ···················· | ···················· | ···················· | ···················· | ···················· | |
| Q | 6161 hCoV-19/Northern Ireland/NIRE-108B33/2020\|EPI_ISL_ | ···················· | ···················· | ···················· | ···················· | ···················· | ···················· | |
| Q | 6162 hCoV-19/Northern Ireland/NIRE-108B42/2020\|EPI_ISL_ | ···················· | ···················· | ···················· | ···················· | ···················· | ···················· | |
| Q | 6163 hCoV-19/Northern Ireland/NIRE-108B51/2020\|EPI_ISL_ | ···················· | ···················· | ···················· | ···················· | ···················· | ···················· | |
| Q | 6164 hCoV-19/Northern Ireland/NIRE-108B8E/2020\|EPI_ISL_ | ···················· | ···················· | ···················· | ···················· | ···················· | ···················· | |
| Q | 6166 hCoV-19/Northern Ireland/NIRE-108BD9/2020\|EPI_ISL_ | ···················· | ···················· | ···················· | ···················· | ···················· | ···················· | |
| Q | 6168 hCoV-19/Northern Ireland/NIRE-108BF7/2020\|EPI_ISL_ | ···················· | ···················· | ···················· | ···················· | ···················· | ···················· | |
| Q | 6169 hCoV-19/Northern Ireland/NIRE-108C03/2020\|EPI_ISL_ | ···················· | ···················· | ···················· | ···················· | ·NNN················ | ···················· | 1 |
| Q | 6170 hCoV-19/Northern Ireland/NIRE-108C12/2020\|EPI_ISL_ | ···················· | ···················· | ···················· | ···················· | ···················· | ···················· | |
| Q | 6171 hCoV-19/Northern Ireland/NIRE-108C5E/2020\|EPI_ISL_ | ···················· | ···················· | ···················· | ···················· | ···················· | ···················· | |
| Q | 6172 hCoV-19/Northern Ireland/NIRE-108C6D/2020\|EPI_ISL_ | ···················· | ···················· | ···················· | ···················· | ···················· | ···················· | |
| Q | 6173 hCoV-19/Northern Ireland/NIRE-108CB8/2020\|EPI_ISL_ | ···················· | ···················· | ···················· | ···················· | ···················· | ···················· | |
| Q | 6174 hCoV-19/Northern Ireland/NIRE-108CC7/2020\|EPI_ISL_ | ···················· | ···················· | ···················· | ···················· | ···················· | ···················· | |

| | | | | | | | |
|---|---|---|---|---|---|---|---|
| Q | 6176 hCoV-19/Northern Ireland/NIRE-108CF4/2020\|EPI_ISL_ | .................... | .................... | .................... | .................... | .................... | .................... |
| Q | 6177 hCoV-19/Northern Ireland/NIRE-108D00/2020\|EPI_ISL_ | .................... | .................... | .................... | .................... | .................... | .................... |
| Q | 6178 hCoV-19/Northern Ireland/NIRE-108D3D/2020\|EPI_ISL_ | .................... | .................... | .................... | .................... | .................... | .................... |
| Q | 6179 hCoV-19/Northern Ireland/NIRE-108D5B/2020\|EPI_ISL_ | .................... | .................... | .................... | .................... | .................... | .................... |
| Q | 6180 hCoV-19/Northern Ireland/NIRE-108D6A/2020\|EPI_ISL_ | .................... | .................... | .................... | .................... | .................... | .................... |
| Q | 6181 hCoV-19/Northern Ireland/NIRE-108D88/2020\|EPI_ISL_ | .................... | .................... | .................... | .................... | .................... | .................... |
| Q | 6182 hCoV-19/Northern Ireland/NIRE-108DA6/2020\|EPI_ISL_ | .................... | .................... | .................... | .................... | .................... | .................... |
| Q | 6183 hCoV-19/Northern Ireland/NIRE-108DB5/2020\|EPI_ISL_ | .................... | .................... | .................... | .................... | .................... | .................... |
| Q | 6184 hCoV-19/Northern Ireland/NIRE-108DC4/2020\|EPI_ISL_ | .................... | .................... | .................... | .................... | .................... | .................... |
| Q | 6185 hCoV-19/Northern Ireland/NIRE-108DD3/2020\|EPI_ISL_ | .................... | .................... | .................... | .................... | .................... | .................... |
| Q | 6186 hCoV-19/Northern Ireland/NIRE-108DE2/2020\|EPI_ISL_ | .................... | .................... | .................... | .................... | .................... | .................... |
| Q | 6187 hCoV-19/Northern Ireland/NIRE-108E0D/2020\|EPI_ISL_ | .................... | .................... | .................... | .................... | .................... | .................... |
| Q | 6188 hCoV-19/Northern Ireland/NIRE-108E2B/2020\|EPI_ISL_ | .................... | .................... | .................... | .................... | .................... | .................... |
| Q | 6190 hCoV-19/Northern Ireland/NIRE-108E58/2020\|EPI_ISL_ | .................... | .................... | .................... | .................... | .................... | .................... |
| Q | 6191 hCoV-19/Northern Ireland/NIRE-108E67/2020\|EPI_ISL_ | .................... | .................... | .................... | .................... | .................... | .................... |
| Q | 6192 hCoV-19/Northern Ireland/NIRE-108E85/2020\|EPI_ISL_ | .................... | .................... | .................... | .................... | .................... | .................... |
| Q | 6193 hCoV-19/England/NORW-F01F1/2020\|EPI_ISL_627570\|202 | .................... | .A.................. | .................... | .................... | .................... | .................... |
| Q | 6194 hCoV-19/England/NORW-F023A/2020\|EPI_ISL_627571\|202 | .................... | .................... | .................... | .................... | .................... | .................... |
| Q | 6196 hCoV-19/England/NORW-F02B2/2020\|EPI_ISL_627573\|202 | .................... | .................... | .................... | .................... | .................... | .................... |
| Q | 6197 hCoV-19/England/NORW-F02C1/2020\|EPI_ISL_627574\|202 | .................... | .................... | .................... | .................... | .................... | .................... |
| Q | 6198 hCoV-19/England/NORW-F02D0/2020\|EPI_ISL_627575\|202 | .................... | .................... | .................... | .................... | .................... | .................... |
| Q | 6199 hCoV-19/England/NORW-F02EF/2020\|EPI_ISL_627576\|202 | .................... | .................... | .................... | .................... | .................... | .................... |
| Q | 6203 hCoV-19/England/NORW-F0346/2020\|EPI_ISL_627580\|202 | .................... | .................... | .................... | .................... | .................... | .................... |
| Q | 6204 hCoV-19/England/NORW-F0355/2020\|EPI_ISL_627581\|202 | .................... | .................... | .................... | .................... | .................... | .................... |
| Q | 6206 hCoV-19/England/NORW-F0382/2020\|EPI_ISL_627583\|202 | .................... | .................... | .................... | .................... | .................... | .................... |
| Q | 6207 hCoV-19/England/NORW-F05D7/2020\|EPI_ISL_627584\|202 | .................... | .................... | .................... | .................... | ......T............. | .................... |
| Q | 6210 hCoV-19/England/NORW-F0F06/2020\|EPI_ISL_627587\|202 | .................... | .................... | .................... | .................... | .................... | .................... |
| Q | 6211 hCoV-19/England/NORW-F0F33/2020\|EPI_ISL_627588\|202 | .................... | .................... | .................... | .................... | .................... | .................... |
| Q | 6212 hCoV-19/England/NORW-F0F60/2020\|EPI_ISL_627589\|202 | .................... | .................... | .................... | .................... | .................... | .................... |
| Q | 6214 hCoV-19/England/NORW-F0F8E/2020\|EPI_ISL_627591\|202 | .................... | .................... | .................... | .................... | .................... | .................... |
| Q | 6216 hCoV-19/England/NORW-F110F/2020\|EPI_ISL_627593\|202 | .................... | .................... | .................... | .................... | .................... | .................... |
| Q | 6217 hCoV-19/England/NORW-F111E/2020\|EPI_ISL_627594\|202 | .................... | .................... | .................... | .................... | .................... | .................... |
| Q | 6219 hCoV-19/England/NORW-F1187/2020\|EPI_ISL_627596\|202 | .................... | .................... | .................... | .................... | .................... | .................... |
| Q | 6220 hCoV-19/England/NORW-F14BB/2020\|EPI_ISL_627597\|202 | .................... | .................... | .................... | .................... | .................... | .................... |
| Q | 6221 hCoV-19/England/NORW-F14D9/2020\|EPI_ISL_627598\|202 | .................... | .................... | .................... | .................... | .................... | .................... |
| Q | 6222 hCoV-19/England/NORW-F14F7/2020\|EPI_ISL_627599\|202 | .................... | .................... | .................... | .................... | .................... | .................... |
| Q | 6223 hCoV-19/USA/MI-UM-10036468985/2020\|EPI_ISL_628804\| | .................... | .................... | .................... | .................... | .................... | .................... |
| Q | 6224 hCoV-19/USA/MI-UM-10036437656/2020\|EPI_ISL_628805\| | .................... | .................... | .................... | .................... | .................... | .................... |
| Q | 6225 hCoV-19/USA/MI-UM-10036518033/2020\|EPI_ISL_628806\| | .................... | .................... | .................... | .................... | .................... | .................... |
| Q | 6226 hCoV-19/USA/MI-UM-10036461122/2020\|EPI_ISL_628807\| | .................... | .................... | .................... | .................... | .................... | .................... |
| Q | 6227 hCoV-19/USA/MI-UM-10036461388/2020\|EPI_ISL_628808\| | .................... | .................... | .................... | .................... | .................... | .................... |
| Q | 6228 hCoV-19/USA/MI-UM-10036484001/2020\|EPI_ISL_628809\| | .................... | .................... | .................... | .................... | .................... | .................... |
| Q | 6230 hCoV-19/USA/MI-UM-10036486626/2020\|EPI_ISL_628811\| | .................... | .................... | .................... | .................... | .................... | .................... |
| Q | 6231 hCoV-19/USA/MI-UM-10036482489/2020\|EPI_ISL_628813\| | .................... | .................... | .................... | .................... | .................... | .................... |
| Q | 6232 hCoV-19/USA/MI-UM-10036453787/2020\|EPI_ISL_628814\| | .................... | .................... | .................... | .................... | .................... | .................... |
| Q | 6235 hCoV-19/USA/MI-UM-10036475782/2020\|EPI_ISL_628817\| | .................... | .............A...... | .................... | .................... | .................... | .................... |
| Q | 6237 hCoV-19/USA/MI-UM-10036466026/2020\|EPI_ISL_628819\| | .................... | .................... | .................... | .................... | .................... | .................... |
| Q | 6238 hCoV-19/USA/MI-UM-10036483429/2020\|EPI_ISL_628820\| | .................... | .............A...... | .................... | .................... | .................... | .................... |
| Q | 6239 hCoV-19/USA/MI-UM-10036483559/2020\|EPI_ISL_628821\| | .................... | .............A...... | .................... | .................... | .................... | .................... |
| Q | 6240 hCoV-19/USA/MI-UM-10036474937/2020\|EPI_ISL_628822\| | .................... | .................... | .................... | .................... | .................... | .................... |
| Q | 6242 hCoV-19/USA/MI-UM-10036478916/2020\|EPI_ISL_628824\| | .................... | .................... | .................... | .................... | .................... | .................... |
| Q | 6243 hCoV-19/USA/MI-UM-10036459813/2020\|EPI_ISL_628826\| | .................... | .................... | .................... | .................... | .................... | .................... |
| Q | 6244 hCoV-19/USA/MI-UM-10036485871/2020\|EPI_ISL_628828\| | .................... | .................... | .................... | .................... | .................... | .................... |
| Q | 6245 hCoV-19/USA/MI-UM-10036476828/2020\|EPI_ISL_628829\| | .................... | .A.................. | .................... | .................... | .................... | .................... |
| Q | 6247 hCoV-19/USA/MI-UM-10036486712/2020\|EPI_ISL_628831\| | .................... | .................... | .................... | .................... | .................... | .................... |
| Q | 6248 hCoV-19/USA/MI-UM-10036462504/2020\|EPI_ISL_628832\| | .................... | .................... | .................... | .................... | .................... | .................... |
| Q | 6249 hCoV-19/USA/MI-UM-10036484282/2020\|EPI_ISL_628833\| | .................... | .A....A............. | .................... | .................... | .................... | .................... |
| Q | 6250 hCoV-19/USA/MI-UM-10036454976/2020\|EPI_ISL_628834\| | .................... | .................... | .................... | .................... | .................... | .................... |
| Q | 6251 hCoV-19/USA/MI-UM-10036456089/2020\|EPI_ISL_628835\| | .................... | .................... | .................... | .................... | .................... | .................... |
| Q | 6252 hCoV-19/USA/MI-UM-10036455942/2020\|EPI_ISL_628836\| | .................... | .................... | .................... | .................... | .................... | .................... |
| Q | 6254 hCoV-19/USA/MI-UM-10036490199/2020\|EPI_ISL_628861\| | .................... | .................... | .................... | .................... | .................... | .................... |
| Q | 6258 hCoV-19/USA/MI-UM-10036515165/2020\|EPI_ISL_628869\| | .................... | .................... | .................... | .................... | .................... | .................... |
| Q | 6261 hCoV-19/USA/MI-UM-10036515267/2020\|EPI_ISL_628874\| | .................... | .................... | .................... | .................... | .................... | .................... |
| Q | 6262 hCoV-19/USA/MI-UM-10036515220/2020\|EPI_ISL_628875\| | .................... | .................... | .................... | .................... | .................... | .................... |
| Q | 6263 hCoV-19/USA/MI-UM-10036507431/2020\|EPI_ISL_628876\| | .................... | .................... | .................... | .................... | .................... | .................... |
| Q | 6264 hCoV-19/USA/MI-UM-10036515334/2020\|EPI_ISL_628879\| | .................... | .................... | .................... | .................... | .................... | .................... |
| Q | 6265 hCoV-19/USA/MI-UM-10036516003/2020\|EPI_ISL_628880\| | .................... | .................... | .................... | .................... | .................... | .................... |
| Q | 6267 hCoV-19/USA/MI-UM-10036524165/2020\|EPI_ISL_628882\| | .................... | ......A............. | .................... | .................... | .................... | .................... |
| Q | 6269 hCoV-19/USA/NY-NYCPHL-000602/2020\|EPI_ISL_632060\|2 | .................... | .................... | .................... | .................... | .................... | .................... |
| Q | 6270 hCoV-19/USA/NY-NYCPHL-000603/2020\|EPI_ISL_632061\|2 | .................... | .................... | .................... | .................... | .................... | .................... |
| Q | 6271 hCoV-19/USA/NY-NYCPHL-000604/2020\|EPI_ISL_632062\|2 | .................... | .................... | .................... | .................... | .................... | .................... |
| Q | 6272 hCoV-19/USA/NY-NYCPHL-000606/2020\|EPI_ISL_632063\|2 | .................... | .................... | .................... | .................... | .................... | .................... |
| Q | 6273 hCoV-19/USA/NY-NYCPHL-000607/2020\|EPI_ISL_632064\|2 | .................... | .................... | .................... | .................... | .................... | .................... |
| Q | 6274 hCoV-19/USA/NY-NYCPHL-000608/2020\|EPI_ISL_632065\|2 | .................... | .................... | .................... | .................... | .................... | .................... |
| Q | 6275 hCoV-19/USA/NY-NYCPHL-000609/2020\|EPI_ISL_632066\|2 | .................... | .................... | .................... | .................... | .................... | .................... |
| Q | 6276 hCoV-19/USA/NY-NYCPHL-000610/2020\|EPI_ISL_632067\|2 | .................... | .................... | .................... | .................... | .................... | .................... |
| Q | 6277 hCoV-19/USA/NY-NYCPHL-000611/2020\|EPI_ISL_632068\|2 | .................... | .................... | .................... | .................... | .................... | .................... |
| Q | 6278 hCoV-19/USA/NY-NYCPHL-000612/2020\|EPI_ISL_632069\|2 | .................... | .................... | .................... | .................... | .................... | .................... |
| Q | 6279 hCoV-19/USA/NY-NYCPHL-000613/2020\|EPI_ISL_632070\|2 | .................... | .................... | .................... | .................... | .................... | .................... |
| Q | 6280 hCoV-19/USA/NY-NYCPHL-000614/2020\|EPI_ISL_632071\|2 | .................... | .................... | .................... | .................... | .................... | .................... |

| | | | | | | | | |
|---|---|---|---|---|---|---|---|---|
| Q | 6281 | hCoV-19/USA/NY-NYCPHL-000615/2020\|EPI_ISL_632072\|2 | .................... | .................... | .................... | .................... | .................... | .................... |
| Q | 6282 | hCoV-19/USA/NY-NYCPHL-000617/2020\|EPI_ISL_632073\|2 | .................... | .................... | .................... | .................... | .................... | .................... |
| Q | 6283 | hCoV-19/USA/NY-NYCPHL-000618/2020\|EPI_ISL_632074\|2 | .................... | .................... | .................... | .................... | .................... | .................... |
| Q | 6284 | hCoV-19/USA/NY-NYCPHL-000619/2020\|EPI_ISL_632075\|2 | .................... | .................... | .................... | .................... | .................... | .................... |
| Q | 6285 | hCoV-19/USA/NY-NYCPHL-000620/2020\|EPI_ISL_632076\|2 | .................... | .................... | .................... | .................... | .................... | .................... |
| Q | 6286 | hCoV-19/USA/NY-NYCPHL-000621/2020\|EPI_ISL_632077\|2 | .................... | .................... | .................... | .................... | .................... | .................... |
| Q | 6287 | hCoV-19/USA/NY-NYCPHL-000622/2020\|EPI_ISL_632078\|2 | .................... | .................... | .................... | .................... | .................... | .................... |
| Q | 6288 | hCoV-19/USA/NY-NYCPHL-000623/2020\|EPI_ISL_632079\|2 | .................... | .................... | .................... | .................... | .................... | .................... |
| Q | 6289 | hCoV-19/USA/NY-NYCPHL-000624/2020\|EPI_ISL_632080\|2 | .................... | .................... | .................... | .................... | .................... | .................... |
| Q | 6290 | hCoV-19/USA/NY-NYCPHL-000673/2020\|EPI_ISL_632091\|2 | .................... | .................... | .................... | .................... | .................... | .................... |
| Q | 6291 | hCoV-19/USA/NY-NYCPHL-000674/2020\|EPI_ISL_632092\|2 | .................... | .................... | .................... | .................... | .................... | .................... |
| Q | 6292 | hCoV-19/USA/NY-NYCPHL-000675/2020\|EPI_ISL_632093\|2 | .................... | .................... | .................... | .................... | .................... | .................... |
| Q | 6293 | hCoV-19/USA/NY-NYCPHL-000676/2020\|EPI_ISL_632094\|2 | .................... | .................... | .................... | .................... | .................... | .................... |
| Q | 6294 | hCoV-19/USA/NY-NYCPHL-000677/2020\|EPI_ISL_632095\|2 | .................... | .................... | .................... | .................... | .................... | .................... |
| Q | 6295 | hCoV-19/USA/NY-NYCPHL-000678/2020\|EPI_ISL_632096\|2 | .................... | .................... | .................... | .................... | .................... | .................... |
| Q | 6296 | hCoV-19/USA/NY-NYCPHL-000679/2020\|EPI_ISL_632097\|2 | .................... | .................... | .................... | .................... | .................... | .................... |
| Q | 6297 | hCoV-19/USA/NY-NYCPHL-000681/2020\|EPI_ISL_632098\|2 | .................... | .................... | .................... | .................... | .................... | .................... |
| Q | 6298 | hCoV-19/USA/NY-NYCPHL-000682/2020\|EPI_ISL_632099\|2 | .................... | .................... | .................... | .................... | .................... | .................... |
| Q | 6299 | hCoV-19/England/CAMC-AEAF9C/2020\|EPI_ISL_633300\|20 | .................... | .................... | .................... | .................... | .................... | ...A................ |
| Q | 6300 | hCoV-19/England/QEUH-AE4CB9/2020\|EPI_ISL_633301\|20 | .................... | .................... | .................... | .................... | .................... | .................... |
| Q | 6301 | hCoV-19/England/CAMC-AEB0F2/2020\|EPI_ISL_633302\|20 | .................... | .................... | .................... | .................... | .................... | .................... |
| Q | 6302 | hCoV-19/England/QEUH-AE4DB6/2020\|EPI_ISL_633303\|20 | .................... | .................... | .................... | .................... | .................... | .................... |
| Q | 6303 | hCoV-19/England/QEUH-AE4FDE/2020\|EPI_ISL_633304\|20 | .................... | .................... | .................... | .................... | .................... | .................... |
| Q | 6304 | hCoV-19/England/CAMC-AEABB6/2020\|EPI_ISL_633390\|20 | .................... | .................... | .................... | .................... | .................... | .................... |
| Q | 6305 | hCoV-19/England/QEUH-AE4A91/2020\|EPI_ISL_633391\|20 | .................... | .................... | .................... | .................... | .................... | .................... |
| Q | 6306 | hCoV-19/England/QEUH-AE50DA/2020\|EPI_ISL_633392\|20 | .................... | .................... | .................... | .................... | .................... | .................... |
| Q | 6307 | hCoV-19/England/CAMC-AEADCF/2020\|EPI_ISL_633393\|20 | .................... | .................... | .................... | .................... | .................... | .................... |
| Q | 6308 | hCoV-19/England/QEUH-AE530E/2020\|EPI_ISL_633394\|20 | .................... | .................... | .................... | .................... | .................... | .................... |
| Q | 6309 | hCoV-19/England/CAMC-AEAA5F/2020\|EPI_ISL_633395\|20 | .................... | .................... | .................... | .................... | .................... | .................... |
| Q | 6310 | hCoV-19/England/QEUH-AE5395/2020\|EPI_ISL_633396\|20 | .................... | .................... | .................... | .................... | .................... | .................... |
| Q | 6311 | hCoV-19/England/CAMC-AEB0B6/2020\|EPI_ISL_633397\|20 | .................... | .................... | .................... | .................... | .................... | .................... |
| Q | 6312 | hCoV-19/England/CAMC-AEA7D0/2020\|EPI_ISL_633398\|20 | .................... | .................... | .................... | .................... | .................... | .................... |
| Q | 6313 | hCoV-19/England/CAMC-AEAB2F/2020\|EPI_ISL_633399\|20 | .................... | .................... | .................... | .................... | .................... | .................... |
| Q | 6314 | hCoV-19/England/QEUH-AD5BAF/2020\|EPI_ISL_634600\|20 | .................... | .................... | .................... | .................... | .................... | .................... |
| Q | 6315 | hCoV-19/England/QEUH-AD196D/2020\|EPI_ISL_634601\|20 | .................... | .................... | .................... | .................... | .................... | .................... |
| Q | 6316 | hCoV-19/England/QEUH-AD4171/2020\|EPI_ISL_634602\|20 | .................... | .................... | .................... | .................... | .................... | .................... |
| Q | 6317 | hCoV-19/England/QEUH-AD16EE/2020\|EPI_ISL_634603\|20 | .................... | .................... | .................... | .................... | .................... | .................... |
| Q | 6318 | hCoV-19/England/QEUH-AD1A4C/2020\|EPI_ISL_634605\|20 | .................... | .................... | .................... | .................... | .................... | .................... |
| Q | 6319 | hCoV-19/England/QEUH-AD1E6E/2020\|EPI_ISL_634606\|20 | .................... | .................... | .................... | .................... | .................... | .................... |
| Q | 6320 | hCoV-19/England/QEUH-AD45C0/2020\|EPI_ISL_634607\|20 | .................... | .................... | .................... | .................... | .................... | .................... |
| Q | 6321 | hCoV-19/England/QEUH-AD5B72/2020\|EPI_ISL_634608\|20 | .................... | .................... | .................... | .................... | .................... | .................... |
| Q | 6322 | hCoV-19/England/QEUH-AD576F/2020\|EPI_ISL_634609\|20 | .................... | .................... | .................... | .................... | .................... | .................... |
| Q | 6323 | hCoV-19/England/QEUH-AD3FA5/2020\|EPI_ISL_634610\|20 | .................... | .................... | .................... | .................... | .................... | .................... |
| Q | 6324 | hCoV-19/England/QEUH-AD0FA8/2020\|EPI_ISL_634611\|20 | .................... | .................... | .................... | .................... | .................... | .................... |
| Q | 6325 | hCoV-19/England/QEUH-AD5B09/2020\|EPI_ISL_634612\|20 | .................... | .................... | .................... | .................... | .................... | .................... |
| Q | 6326 | hCoV-19/England/QEUH-AD3F69/2020\|EPI_ISL_634614\|20 | .................... | .................... | .................... | .................... | ....T............... | .................... |
| Q | 6327 | hCoV-19/England/QEUH-AD588A/2020\|EPI_ISL_634615\|20 | .................... | .................... | .................... | .................... | .................... | .................... |
| Q | 6328 | hCoV-19/England/QEUH-AD3FC3/2020\|EPI_ISL_634616\|20 | .................... | .................... | .................... | .................... | .................... | .................... |
| Q | 6329 | hCoV-19/England/QEUH-AD5E88/2020\|EPI_ISL_634617\|20 | .................... | .................... | .................... | .................... | .................... | .................... |
| Q | 6330 | hCoV-19/England/QEUH-AD1ED7/2020\|EPI_ISL_634618\|20 | .................... | .................... | .................... | .................... | .................... | .................... |
| Q | 6331 | hCoV-19/England/QEUH-AD4223/2020\|EPI_ISL_634619\|20 | .................... | .................... | .................... | .................... | .................... | .................... |
| Q | 6332 | hCoV-19/England/QEUH-AD5ADF/2020\|EPI_ISL_634620\|20 | .................... | .................... | .................... | .................... | .................... | .................... |
| Q | 6333 | hCoV-19/England/QEUH-AD5468/2020\|EPI_ISL_634621\|20 | .................... | .................... | .................... | .................... | .................... | .................... |
| Q | 6334 | hCoV-19/England/QEUH-AD41AE/2020\|EPI_ISL_634622\|20 | .................... | .................... | .................... | .................... | .................... | .................... |
| Q | 6335 | hCoV-19/England/QEUH-AD1A5B/2020\|EPI_ISL_634623\|20 | .................... | .A.................. | .................... | .................... | .................... | .................... |
| Q | 6336 | hCoV-19/England/QEUH-AD5E5B/2020\|EPI_ISL_634624\|20 | .................... | .................... | .................... | .................... | .................... | .................... |
| Q | 6337 | hCoV-19/England/QEUH-AD4609/2020\|EPI_ISL_634625\|20 | .................... | .................... | .................... | .................... | .................... | .................... |
| Q | 6338 | hCoV-19/England/QEUH-AD58D5/2020\|EPI_ISL_634626\|20 | .................... | .................... | .................... | .................... | .................... | .................... |
| Q | 6339 | hCoV-19/England/QEUH-AD4654/2020\|EPI_ISL_634627\|20 | .................... | .................... | .................... | .................... | .................... | .................... |
| Q | 6340 | hCoV-19/England/QEUH-AD1693/2020\|EPI_ISL_634628\|20 | .................... | .................... | .................... | .................... | .................... | .................... |
| Q | 6341 | hCoV-19/England/QEUH-AD5A1B/2020\|EPI_ISL_634629\|20 | .................... | .................... | .................... | .................... | .................... | .................... |
| Q | 6342 | hCoV-19/England/QEUH-AD4487/2020\|EPI_ISL_634630\|20 | .................... | .................... | .................... | .................... | .................... | .................... |
| Q | 6343 | hCoV-19/England/QEUH-AD41DB/2020\|EPI_ISL_634631\|20 | .................... | .................... | .................... | .................... | .................... | .................... |
| Q | 6344 | hCoV-19/England/QEUH-AD4690/2020\|EPI_ISL_634632\|20 | .................... | .................... | .................... | .................... | .................... | .................... |
| Q | 6345 | hCoV-19/England/QEUH-AD1718/2020\|EPI_ISL_634633\|20 | .................... | .................... | .................... | .................... | .................... | .................... |
| Q | 6346 | hCoV-19/England/QEUH-AD19F4/2020\|EPI_ISL_634634\|20 | .................... | .................... | .................... | .................... | .................... | .................... |
| Q | 6347 | hCoV-19/England/QEUH-AD4BFB/2020\|EPI_ISL_634635\|20 | .................... | .................... | .................... | .................... | .................... | .................... |
| Q | 6348 | hCoV-19/USA/CA-ALSR-3174/2020\|EPI_ISL_635900\|2020- | .................... | .................... | .................... | .................... | .................... | .................... |
| Q | 6349 | hCoV-19/USA/CA-ALSR-3188/2020\|EPI_ISL_635905\|2020- | .................... | .A.................. | .................... | .................... | .................... | .................... |
| Q | 6350 | hCoV-19/USA/CA-ALSR-3190/2020\|EPI_ISL_635906\|2020- | .................... | .A.................. | .................... | .................... | .................... | .................... |
| Q | 6351 | hCoV-19/USA/CA-ALSR-3193/2020\|EPI_ISL_635907\|2020- | .................... | .A.................. | .................... | .................... | .................... | .................... |
| Q | 6352 | hCoV-19/USA/CA-ALSR-3196/2020\|EPI_ISL_635908\|2020- | .................... | .A.................. | .................... | .................... | .................... | .................... |
| Q | 6353 | hCoV-19/USA/CA-ALSR-3210/2020\|EPI_ISL_635912\|2020- | .................... | .................... | .................... | .................... | .................... | .................... |
| Q | 6354 | hCoV-19/USA/CA-ALSR-3212/2020\|EPI_ISL_635913\|2020- | .................... | .................... | .................... | .................... | .................... | .................... |
| Q | 6355 | hCoV-19/USA/CA-ALSR-3214/2020\|EPI_ISL_635914\|2020- | .................... | .................... | .................... | .................... | .................... | .................... |
| Q | 6356 | hCoV-19/USA/CA-ALSR-3217/2020\|EPI_ISL_635917\|2020- | .................... | .A.................. | .................... | .................... | .................... | .................... |
| Q | 6357 | hCoV-19/USA/CA-ALSR-3218/2020\|EPI_ISL_635918\|2020- | .................... | .................... | .................... | .................... | .................... | .................... |
| Q | 6358 | hCoV-19/USA/CA-ALSR-3219/2020\|EPI_ISL_635919\|2020- | .................... | .................... | .................... | .................... | .................... | .................... |
| Q | 6359 | hCoV-19/USA/CA-ALSR-3221/2020\|EPI_ISL_635920\|2020- | .................... | .................... | .................... | .................... | .................... | .................... |
| Q | 6360 | hCoV-19/USA/CA-ALSR-3222/2020\|EPI_ISL_635921\|2020- | .................... | .................... | .................... | .................... | .................... | .................... |

Q 6361 hCoV-19/USA/CA-ALSR-3224/2020|EPI_ISL_635922|2020- ···················· ·A·················· ···················· ···················· ···················· ····················
Q 6362 hCoV-19/USA/CA-ALSR-3226/2020|EPI_ISL_635923|2020- ···················· ···················· ···················· ···················· ···················· ····················
Q 6363 hCoV-19/USA/CA-ALSR-3228/2020|EPI_ISL_635924|2020- ···················· ·A·················· ···················· ···················· ···················· ····················
Q 6364 hCoV-19/USA/CA-ALSR-3229/2020|EPI_ISL_635925|2020- ···················· ·A·················· ···················· ···················· ···················· ····················
Q 6365 hCoV-19/USA/CA-ALSR-3239/2020|EPI_ISL_635929|2020- ···················· ·A·················· ···················· ···················· ···················· ····················
Q 6366 hCoV-19/USA/CA-ALSR-3241/2020|EPI_ISL_635930|2020- ···················· ···················· ···················· ···················· ···················· ····················
Q 6367 hCoV-19/USA/CA-ALSR-3243/2020|EPI_ISL_635931|2020- ···················· ·A·················· ···················· ···················· ···················· ····················
Q 6368 hCoV-19/USA/CA-ALSR-3256/2020|EPI_ISL_635932|2020- ···················· ·A·················· ···················· ···················· ···················· ····················
Q 6369 hCoV-19/USA/CA-ALSR-3257/2020|EPI_ISL_635933|2020- ···················· ·A·················· ···················· ···················· ···················· ····················
Q 6370 hCoV-19/USA/CA-ALSR-3258/2020|EPI_ISL_635934|2020- ···················· ·A·················· ···················· ···················· ···················· ····················
Q 6371 hCoV-19/USA/CA-ALSR-3259/2020|EPI_ISL_635935|2020- ···················· ·A·················· ···················· ···················· ···················· ····················
Q 6372 hCoV-19/USA/CA-ALSR-3260/2020|EPI_ISL_635936|2020- ···················· ·A·················· ···················· ···················· ···················· ····················
Q 6373 hCoV-19/USA/CA-ALSR-3262/2020|EPI_ISL_635937|2020- ···················· ·A·················· ···················· ···················· ···················· ····················
Q 6374 hCoV-19/USA/CA-ALSR-3264/2020|EPI_ISL_635939|2020- ···················· ·A·················· ···················· ···················· ···················· ····················
Q 6375 hCoV-19/USA/CA-ALSR-3265/2020|EPI_ISL_635940|2020- ···················· ·A·················· ···················· ···················· ···················· ····················
Q 6376 hCoV-19/USA/CA-ALSR-3266/2020|EPI_ISL_635941|2020- ···················· ·A·················· ···················· ···················· ···················· ····················
Q 6377 hCoV-19/USA/CA-ALSR-3267/2020|EPI_ISL_635942|2020- ···················· ·A·················· ···················· ···················· ···················· ····················
Q 6378 hCoV-19/USA/CA-ALSR-3268/2020|EPI_ISL_635943|2020- ···················· ·A·················· ···················· ···················· ···················· ····················
Q 6379 hCoV-19/USA/CA-ALSR-3269/2020|EPI_ISL_635944|2020- ···················· ·A·················· ···················· ···················· ···················· ····················
Q 6380 hCoV-19/USA/CA-ALSR-3270/2020|EPI_ISL_635945|2020- ···················· ·A·················· ···················· ···················· ···················· ····················
Q 6381 hCoV-19/USA/CA-ALSR-3273/2020|EPI_ISL_635946|2020- ···················· ·A·················· ···················· ···················· ···················· ····················
Q 6382 hCoV-19/USA/CA-ALSR-3277/2020|EPI_ISL_635947|2020- ···················· ·A·················· ···················· ···················· ···················· ····················
Q 6383 hCoV-19/USA/CA-ALSR-3281/2020|EPI_ISL_635948|2020- ···················· ·A·················· ···················· ···················· ···················· ····················
Q 6384 hCoV-19/USA/CA-ALSR-3283/2020|EPI_ISL_635949|2020- ···················· ·A·················· ···················· ···················· ···················· ····················
Q 6385 hCoV-19/USA/CA-ALSR-3287/2020|EPI_ISL_635950|2020- ···················· ·A·················· ···················· ···················· ···················· ····················
Q 6386 hCoV-19/USA/CA-ALSR-3289/2020|EPI_ISL_635951|2020- ···················· ···················· ···················· ···················· ···················· ····················
Q 6387 hCoV-19/USA/CA-ALSR-3290/2020|EPI_ISL_635952|2020- ···················· ···················· ···················· ···················· ···················· ····················
Q 6388 hCoV-19/USA/CA-ALSR-3293/2020|EPI_ISL_635953|2020- ···················· ···················· ···················· ···················· ···················· ····················
Q 6389 hCoV-19/USA/CA-ALSR-3304/2020|EPI_ISL_635954|2020- ···················· ·A·················· ···················· ···················· ···················· ····················
Q 6390 hCoV-19/USA/CA-ALSR-3310/2020|EPI_ISL_635956|2020- ···················· ·A·················· ···················· ···················· ···················· ····················
Q 6391 hCoV-19/USA/CA-ALSR-3314/2020|EPI_ISL_635957|2020- ···················· ·A·················· ···················· ···················· ···················· ····················
Q 6392 hCoV-19/USA/CA-ALSR-3316/2020|EPI_ISL_635958|2020- ···················· ·A·················· ···················· ···················· ···················· ····················
Q 6393 hCoV-19/USA/CA-ALSR-3317/2020|EPI_ISL_635959|2020- ···················· ·A·················· ···················· ···················· ···················· ····················
Q 6394 hCoV-19/USA/CA-ALSR-3320/2020|EPI_ISL_635960|2020- ···················· ···················· ···················· ···················· ··A················· ····················
Q 6395 hCoV-19/USA/CA-ALSR-3321/2020|EPI_ISL_635961|2020- ···················· ·A·················· ···················· ···················· ···················· ····················
Q 6396 hCoV-19/USA/CA-ALSR-3322/2020|EPI_ISL_635962|2020- ···················· ···················· ···················· ···················· ···················· ····················
Q 6397 hCoV-19/USA/CA-ALSR-3323/2020|EPI_ISL_635963|2020- ···················· ·A·················· ···················· ···················· ···················· ····················
Q 6398 hCoV-19/USA/CA-ALSR-3324/2020|EPI_ISL_635964|2020- ···················· ·A·················· ···················· ···················· ···················· ····················
Q 6399 hCoV-19/USA/CA-ALSR-3326/2020|EPI_ISL_635965|2020- ···················· ·A·················· ···················· ···················· ···················· ····················
Q 6400 hCoV-19/USA/CA-ALSR-3330/2020|EPI_ISL_635966|2020- ···················· ·A·················· ···················· ···················· ···················· ····················
Q 6401 hCoV-19/USA/NY-NYCPHL-000540/2020|EPI_ISL_632000|2 ···················· ···················· ···················· ···················· ···················· ····················
Q 6402 hCoV-19/USA/NY-NYCPHL-000541/2020|EPI_ISL_632001|2 ···················· ···················· ···················· ···················· ···················· ····················
Q 6403 hCoV-19/USA/NY-NYCPHL-000542/2020|EPI_ISL_632002|2 ···················· ···················· ···················· ···················· ···················· ····················
Q 6404 hCoV-19/USA/NY-NYCPHL-000543/2020|EPI_ISL_632003|2 ···················· ···················· ···················· ···················· ···················· ····················
Q 6405 hCoV-19/USA/NY-NYCPHL-000545/2020|EPI_ISL_632005|2 ···················· ···················· ···················· ···················· ···················· ····················
Q 6406 hCoV-19/USA/NY-NYCPHL-000546/2020|EPI_ISL_632006|2 ···················· ···················· ···················· ···················· ···T················ ····················
Q 6407 hCoV-19/USA/NY-NYCPHL-000547/2020|EPI_ISL_632007|2 ···················· ···················· ···················· ···················· ···················· ····················
Q 6408 hCoV-19/USA/NY-NYCPHL-000548/2020|EPI_ISL_632008|2 ···················· ···················· ···················· ···················· ···················· ····················
Q 6409 hCoV-19/USA/NY-NYCPHL-000549/2020|EPI_ISL_632009|2 ···················· ···················· ···················· ···················· ···················· ····················
Q 6410 hCoV-19/USA/NY-NYCPHL-000550/2020|EPI_ISL_632010|2 ···················· ···················· ···················· ···················· ···················· ····················
Q 6411 hCoV-19/USA/NY-NYCPHL-000551/2020|EPI_ISL_632011|2 ···················· ···················· ···················· ···················· ···················· ····················
Q 6412 hCoV-19/USA/NY-NYCPHL-000552/2020|EPI_ISL_632012|2 ···················· ···················· ···················· ···················· ···················· ····················
Q 6413 hCoV-19/USA/NY-NYCPHL-000553/2020|EPI_ISL_632013|2 ···················· ···················· ···················· ···················· ···················· ····················
Q 6414 hCoV-19/USA/NY-NYCPHL-000554/2020|EPI_ISL_632014|2 ···················· ···················· ···················· ···················· ···················· ····················
Q 6415 hCoV-19/USA/NY-NYCPHL-000556/2020|EPI_ISL_632015|2 ···················· ···················· ···················· ···················· ···················· ····················
Q 6416 hCoV-19/USA/NY-NYCPHL-000557/2020|EPI_ISL_632016|2 ···················· ···················· ···················· ···················· ···················· ····················
Q 6417 hCoV-19/USA/NY-NYCPHL-000558/2020|EPI_ISL_632017|2 ···················· ···················· ···················· ···················· ···················· ····················
Q 6418 hCoV-19/USA/NY-NYCPHL-000559/2020|EPI_ISL_632018|2 ···················· ···················· ···················· ···················· ···················· ····················
Q 6419 hCoV-19/USA/NY-NYCPHL-000560/2020|EPI_ISL_632019|2 ···················· ···················· ···················· ···················· ···················· ····················
Q 6420 hCoV-19/USA/NY-NYCPHL-000561/2020|EPI_ISL_632020|2 ···················· ···················· ···················· ···················· ···················· ····················
Q 6421 hCoV-19/USA/NY-NYCPHL-000562/2020|EPI_ISL_632021|2 ···················· ···················· ···················· ···················· ···················· ····················
Q 6422 hCoV-19/USA/NY-NYCPHL-000563/2020|EPI_ISL_632022|2 ···················· ···················· ···················· ···················· ···················· ····················
Q 6423 hCoV-19/USA/NY-NYCPHL-000564/2020|EPI_ISL_632023|2 ···················· ···················· ···················· ···················· ···················· ····················
Q 6424 hCoV-19/USA/NY-NYCPHL-000565/2020|EPI_ISL_632024|2 ···················· ···················· ···················· ···················· ···················· ····················
Q 6425 hCoV-19/USA/NY-NYCPHL-000566/2020|EPI_ISL_632025|2 ···················· ···················· ···················· ···················· ···················· ····················
Q 6426 hCoV-19/USA/NY-NYCPHL-000567/2020|EPI_ISL_632026|2 ···················· ···················· ···················· ···················· ···················· ····················
Q 6427 hCoV-19/USA/NY-NYCPHL-000568/2020|EPI_ISL_632027|2 ···················· ···················· ···················· ···················· ···················· ····················
Q 6428 hCoV-19/USA/NY-NYCPHL-000569/2020|EPI_ISL_632028|2 ···················· ···················· ···················· ···················· ···················· ····················
Q 6429 hCoV-19/USA/NY-NYCPHL-000570/2020|EPI_ISL_632029|2 ···················· ···················· ···················· ···················· ···················· ····················
Q 6430 hCoV-19/USA/NY-NYCPHL-000571/2020|EPI_ISL_632030|2 ···················· ···················· ···················· ···················· ···················· ····················
Q 6431 hCoV-19/USA/NY-NYCPHL-000572/2020|EPI_ISL_632031|2 ···················· ···················· ···················· ···················· ···················· ····················
Q 6432 hCoV-19/USA/NY-NYCPHL-000573/2020|EPI_ISL_632032|2 ···················· ···················· ···················· ···················· ···················· ····················
Q 6433 hCoV-19/USA/NY-NYCPHL-000575/2020|EPI_ISL_632034|2 ···················· ···················· ···················· ···················· ···················· ····················
Q 6434 hCoV-19/USA/NY-NYCPHL-000576/2020|EPI_ISL_632035|2 ···················· ···················· ···················· ···················· ···················· ····················
Q 6435 hCoV-19/USA/NY-NYCPHL-000577/2020|EPI_ISL_632036|2 ···················· ···················· ···················· ···················· ···················· ····················
Q 6436 hCoV-19/USA/NY-NYCPHL-000578/2020|EPI_ISL_632037|2 ···················· ···················· ···················· ···················· ···················· ····················
Q 6437 hCoV-19/USA/NY-NYCPHL-000580/2020|EPI_ISL_632038|2 ···················· ···················· ···················· ···················· ···················· ····················
Q 6438 hCoV-19/USA/NY-NYCPHL-000581/2020|EPI_ISL_632039|2 ···················· ···················· ···················· ···················· ···················· ····················
Q 6439 hCoV-19/USA/NY-NYCPHL-000582/2020|EPI_ISL_632040|2 ···················· ···················· ···················· ···················· ···················· ····················
Q 6440 hCoV-19/USA/NY-NYCPHL-000583/2020|EPI_ISL_632041|2 ···················· ···················· ···················· ···················· ···················· ····················

Q 6441 hCoV-19/USA/NY-NYCPHL-000584/2020|EPI_ISL_632042|2 .................... .................... ..................... .................... .................... ....................
Q 6442 hCoV-19/USA/NY-NYCPHL-000585/2020|EPI_ISL_632043|2 .................... .................... ..................... .................... .................... ....................
Q 6443 hCoV-19/USA/NY-NYCPHL-000586/2020|EPI_ISL_632044|2 .................... .................... ..................... .................... .................... ....................
Q 6444 hCoV-19/USA/NY-NYCPHL-000587/2020|EPI_ISL_632045|2 .................... .................... ..................... .................... .................... ....................
Q 6445 hCoV-19/USA/NY-NYCPHL-000588/2020|EPI_ISL_632046|2 .................... .................... ..................... .................... .................... ....................
Q 6446 hCoV-19/USA/NY-NYCPHL-000589/2020|EPI_ISL_632047|2 .................... .................... ..................... .................... .................... ....................
Q 6447 hCoV-19/USA/NY-NYCPHL-000590/2020|EPI_ISL_632048|2 .................... .................... ..................... .................... .................... ....................
Q 6448 hCoV-19/USA/NY-NYCPHL-000591/2020|EPI_ISL_632049|2 .................... .................... ..................... .................... .................... ....................
Q 6449 hCoV-19/USA/NY-NYCPHL-000592/2020|EPI_ISL_632050|2 .................... .................... ..................... .................... .................... ....................
Q 6450 hCoV-19/USA/NY-NYCPHL-000593/2020|EPI_ISL_632051|2 .................... .................... ..................... .................... .................... ....................
Q 6451 hCoV-19/USA/NY-NYCPHL-000594/2020|EPI_ISL_632052|2 .................... .................... ..................... .................... .................... ....................
Q 6452 hCoV-19/USA/NY-NYCPHL-000595/2020|EPI_ISL_632053|2 .................... .................... ..................... .................... .................... ....................
Q 6453 hCoV-19/USA/NY-NYCPHL-000596/2020|EPI_ISL_632054|2 .................... .................... ..................... .................... .................... ....................
Q 6454 hCoV-19/USA/NY-NYCPHL-000597/2020|EPI_ISL_632055|2 .................... .................... ..................... .................... .................... ....................
Q 6455 hCoV-19/USA/NY-NYCPHL-000598/2020|EPI_ISL_632056|2 .................... .................... ..................... .................... .................... ....................
Q 6456 hCoV-19/USA/NY-NYCPHL-000599/2020|EPI_ISL_632057|2 .................... .................... ..................... .................... .................... ....................
Q 6457 hCoV-19/USA/NY-NYCPHL-000600/2020|EPI_ISL_632058|2 .................... .................... ..................... .................... .................... ....................
Q 6458 hCoV-19/USA/NY-NYCPHL-000601/2020|EPI_ISL_632059|2 .................... .................... ..................... .................... .................... ....................
Q 6459 hCoV-19/England/QEUH-AE53E0/2020|EPI_ISL_633305|20 .................... .................... ..................... .................... .................... ....................
Q 6460 hCoV-19/England/QEUH-AE4985/2020|EPI_ISL_633306|20 .................... .................... ..................... .................... .................... ....................
Q 6461 hCoV-19/England/QEUH-AE4C6E/2020|EPI_ISL_633307|20 .................... .................... ..................... .................... .................... ....................
Q 6462 hCoV-19/England/CAMC-AEAD29/2020|EPI_ISL_633308|20 .................... .................... ..................... .................... .................... ....................
Q 6463 hCoV-19/England/QEUH-AE4CE6/2020|EPI_ISL_633309|20 .................... .................... ..................... .................... .................... ....................
Q 6464 hCoV-19/England/QEUH-AE5061/2020|EPI_ISL_633310|20 .................... .................... ..................... .................... .................... ....................
Q 6465 hCoV-19/England/CAMC-AEA970/2020|EPI_ISL_633311|20 .................... .................... ..................... .................... ......T............. ....................
Q 6466 hCoV-19/England/QEUH-AE5368/2020|EPI_ISL_633312|20 .................... .................... ..................... .................... .................... ....................
Q 6467 hCoV-19/England/CAMC-AEAFE7/2020|EPI_ISL_633313|20 .................... .................... ..................... .................... .................... ....................
Q 6468 hCoV-19/England/CAMC-AEAA31/2020|EPI_ISL_633314|20 .................... .................... ..................... .................... .................... ....................
Q 6469 hCoV-19/England/CAMC-AEA70D/2020|EPI_ISL_633315|20 .................... .................... ..................... .................... .................... ....................
Q 6470 hCoV-19/England/QEUH-AE4A55/2020|EPI_ISL_633316|20 .................... .................... ..................... .................... .................... ....................
Q 6471 hCoV-19/England/QEUH-AE5131/2020|EPI_ISL_633317|20 .................... .................... ..................... .................... .................... ....................
Q 6472 hCoV-19/England/QEUH-AE4ADD/2020|EPI_ISL_633318|20 .................... .................... ..................... .................... .................... ....................
Q 6473 hCoV-19/England/CAMC-AEB1C2/2020|EPI_ISL_633319|20 .................... .................... ..................... .................... .................... ....................
Q 6474 hCoV-19/England/QEUH-AE5140/2020|EPI_ISL_633320|20 .................... .................... ..................... .................... ......T............. ....................
Q 6475 hCoV-19/England/CAMC-AEAE53/2020|EPI_ISL_633321|20 .................... .................... ..................... .................... .................... ....................
Q 6476 hCoV-19/England/CAMC-AEAD56/2020|EPI_ISL_633322|20 .................... .................... ..................... .................... ......T............. ....................
Q 6477 hCoV-19/England/QEUH-AE518C/2020|EPI_ISL_633323|20 .................... .................... ..................... .................... .................... ....................
Q 6478 hCoV-19/England/CAMC-AEA7C1/2020|EPI_ISL_633324|20 .................... .................... ..................... .................... .................... ....................
Q 6479 hCoV-19/England/CAMC-AEB07A/2020|EPI_ISL_633325|20 .................... .................... ..................... .................... .................... ....................
Q 6480 hCoV-19/England/QEUH-AE4DE3/2020|EPI_ISL_633326|20 .................... .................... ..................... .................... .................... ....................
Q 6481 hCoV-19/England/QEUH-AE52B6/2020|EPI_ISL_633327|20 .................... .................... ..................... .................... .................... ....................
Q 6482 hCoV-19/England/QEUH-AE53A4/2020|EPI_ISL_633328|20 .................... .................... ..................... .................... .................... ....................
Q 6483 hCoV-19/England/CAMC-AEAAE6/2020|EPI_ISL_633329|20 .................... .................... ..................... .................... .................... ....................
Q 6484 hCoV-19/England/CAMC-AEACB3/2020|EPI_ISL_633330|20 .................... .................... ..................... .................... .................... ....................
Q 6485 hCoV-19/England/QEUH-AE4A46/2020|EPI_ISL_633332|20 .................... .................... ..................... .................... .................... ....................
Q 6486 hCoV-19/England/CAMC-AEAE9F/2020|EPI_ISL_633333|20 .................... .................... ..................... .................... ......T............. ....................
Q 6487 hCoV-19/England/CAMC-AEAAAA/2020|EPI_ISL_633334|20 .................... .................... ..................... .................... .................... ....................
Q 6488 hCoV-19/England/QEUH-AE4994/2020|EPI_ISL_633335|20 .................... .................... ..................... .................... .................... ....................
Q 6489 hCoV-19/England/CAMC-AEA864/2020|EPI_ISL_633336|20 .................... .A.................. ..................... .................... .................... ..A.................
Q 6490 hCoV-19/England/CAMC-AEB089/2020|EPI_ISL_633337|20 .................... .................... ..................... .................... .................... ....................
Q 6491 hCoV-19/England/QEUH-AE533B/2020|EPI_ISL_633338|20 .................... .................... ..................... .................... .................... ....................
Q 6492 hCoV-19/England/QEUH-AE51C8/2020|EPI_ISL_633339|20 .................... .................... ..................... .................... .................... ....................
Q 6493 hCoV-19/England/CAMC-AEB177/2020|EPI_ISL_633340|20 .................... .................... ..................... .................... .................... ....................
Q 6494 hCoV-19/England/QEUH-AE5359/2020|EPI_ISL_633342|20 .................... ........T........... ..................... .................... .................... ....................
Q 6495 hCoV-19/England/QEUH-AE523E/2020|EPI_ISL_633343|20 .................... .................... ..................... .................... .................... ....................
Q 6496 hCoV-19/England/CAMC-AEA64C/2020|EPI_ISL_633344|20 .................... .................... ..................... .................... .................... ....................
Q 6497 hCoV-19/England/CAMC-AEAE71/2020|EPI_ISL_633345|20 .................... .................... ..................... .................... .................... ....................
Q 6498 hCoV-19/England/CAMC-AEA8BF/2020|EPI_ISL_633346|20 .................... .................... ..................... .................... ......T............. ....................
Q 6499 hCoV-19/England/CAMC-AEB1E0/2020|EPI_ISL_633347|20 .................... .................... ..................... .................... ....T............... ....................
Q 6500 hCoV-19/England/QEUH-AE5429/2020|EPI_ISL_633348|20 .................... .................... ..................... .................... .................... ....................
Q 6501 hCoV-19/England/CAMC-AEB0A7/2020|EPI_ISL_633349|20 .................... .................... ..................... .................... .................... ....................
Q 6502 hCoV-19/England/QEUH-AE51AA/2020|EPI_ISL_633351|20 .................... .................... ..................... .................... .................... ....................
Q 6503 hCoV-19/England/QEUH-AE4C13/2020|EPI_ISL_633352|20 .................... .................... ..................... .................... .................... ....................
Q 6504 hCoV-19/England/QEUH-AE516E/2020|EPI_ISL_633353|20 .................... .................... ..................... .................... .................... ....................
Q 6505 hCoV-19/England/QEUH-AE50BC/2020|EPI_ISL_633354|20 .................... .................... ..................... .................... .................... ....................
Q 6506 hCoV-19/England/QEUH-AE4EA4/2020|EPI_ISL_633355|20 .................... .................... ..................... .................... .................... ....................
Q 6507 hCoV-19/England/CAMC-AEA80A/2020|EPI_ISL_633356|20 .................... .................... ..................... .................... .................... ....................
Q 6508 hCoV-19/England/CAMC-AEAD92/2020|EPI_ISL_633357|20 .................... .................... ..................... .................... .................... ....................
Q 6509 hCoV-19/England/QEUH-AE5007/2020|EPI_ISL_633358|20 .................... .................... ..................... .................... .................... ....................
Q 6510 hCoV-19/England/CAMC-AEB13B/2020|EPI_ISL_633359|20 .................... .................... ..................... .................... .................... ....................
Q 6511 hCoV-19/England/CAMC-AEAF6F/2020|EPI_ISL_633360|20 .................... .................... ..................... .................... .................... ....................
Q 6512 hCoV-19/England/QEUH-AE51E6/2020|EPI_ISL_633362|20 .................... .A.................. ..................... .................... .................... ..A.................
Q 6513 hCoV-19/England/CAMC-AEA749/2020|EPI_ISL_633363|20 .................... .................... ..................... .................... .................... ....................
Q 6514 hCoV-19/England/QEUH-AE4FCF/2020|EPI_ISL_633364|20 .................... .................... ..................... .................... .................... ....................
Q 6515 hCoV-19/England/CAMC-AEAE44/2020|EPI_ISL_633365|20 .................... .................... ..................... .................... ......T............. ....................
Q 6516 hCoV-19/England/CAMC-AEABE3/2020|EPI_ISL_633366|20 .................... .................... ..................... .................... .................... ....................
Q 6517 hCoV-19/England/QEUH-AE4E3B/2020|EPI_ISL_633367|20 .................... .................... ..................... .................... .................... ....................
Q 6518 hCoV-19/England/CAMC-AEAA6E/2020|EPI_ISL_633369|20 .................... .................... ..................... .................... .................... ....................
Q 6519 hCoV-19/England/CAMC-AEA767/2020|EPI_ISL_633370|20 .................... .................... ..................... .................... .................... ....................
Q 6520 hCoV-19/England/QEUH-AE4F65/2020|EPI_ISL_633371|20 .................... .................... ..................... .................... .................... ....................

| | | | | | | | |
|---|---|---|---|---|---|---|---|
| Q | 6521 hCoV-19/England/CAMC-AEAC1D/2020\|EPI_ISL_633372\|20 | ···················· | ···················· | ····················· | ···················· | ···················· | ···················· |
| Q | 6522 hCoV-19/England/CAMC-AEACD1/2020\|EPI_ISL_633373\|20 | ···················· | ···················· | ····················· | ···················· | ···················· | ···················· |
| Q | 6523 hCoV-19/England/CAMC-AEAC3B/2020\|EPI_ISL_633374\|20 | ···················· | ···················· | ····················· | ···················· | ···················· | ···················· |
| Q | 6524 hCoV-19/England/CAMC-AEAE17/2020\|EPI_ISL_633375\|20 | ···················· | ···················· | ····················· | ···················· | ···················· | ···················· |
| Q | 6525 hCoV-19/England/QEUH-AE4C22/2020\|EPI_ISL_633376\|20 | ···················· | ···················· | ····················· | ···················· | ···················· | ···················· |
| Q | 6526 hCoV-19/England/QEUH-AE4E1D/2020\|EPI_ISL_633377\|20 | ···················· | ···················· | ····················· | ···················· | ···················· | ···················· |
| Q | 6527 hCoV-19/England/CAMC-AEAAC8/2020\|EPI_ISL_633378\|20 | ···················· | ···················· | ····················· | ···················· | ···················· | ···················· |
| Q | 6528 hCoV-19/England/QEUH-AE4FFC/2020\|EPI_ISL_633379\|20 | ···················· | ···················· | ····················· | ···················· | ···················· | ···················· |
| Q | 6529 hCoV-19/England/CAMC-AEAF7E/2020\|EPI_ISL_633380\|20 | ···················· | ···················· | ····················· | ···················· | ···················· | ···················· |
| Q | 6530 hCoV-19/England/QEUH-AE53D1/2020\|EPI_ISL_633381\|20 | ···················· | ···················· | ····················· | ···················· | ···················· | ···················· |
| Q | 6531 hCoV-19/England/QEUH-AE522F/2020\|EPI_ISL_633382\|20 | ···················· | ···················· | ····················· | ···················· | ······T············· | ···················· |
| Q | 6532 hCoV-19/England/CAMC-AEA907/2020\|EPI_ISL_633383\|20 | ···················· | ···················· | ····················· | ···················· | ······T············· | ···················· |
| Q | 6533 hCoV-19/England/QEUH-AE4E77/2020\|EPI_ISL_633384\|20 | ···················· | ···················· | ····················· | ···················· | ···················· | ···················· |
| Q | 6534 hCoV-19/England/CAMC-AEA846/2020\|EPI_ISL_633385\|20 | ···················· | ···················· | ····················· | ···················· | ···················· | ···················· |
| Q | 6535 hCoV-19/England/QEUH-AE527A/2020\|EPI_ISL_633386\|20 | ···················· | ···················· | ····················· | ···················· | ···················· | ···················· |
| Q | 6536 hCoV-19/England/QEUH-AE531D/2020\|EPI_ISL_633387\|20 | ···················· | ···················· | ····················· | ···················· | ······T············· | ···················· |
| Q | 6537 hCoV-19/England/CAMC-AEB03E/2020\|EPI_ISL_633388\|20 | ···················· | ···················· | ····················· | ···················· | ···················· | ···················· |
| Q | 6538 hCoV-19/England/CAMC-AEA65B/2020\|EPI_ISL_633389\|20 | ···················· | ···················· | ····················· | ···················· | ···················· | ···················· |
| Q | 6539 hCoV-19/England/QEUH-AD104A/2020\|EPI_ISL_634636\|20 | ···················· | ···················· | ····················· | ···················· | ···················· | ···················· |
| Q | 6540 hCoV-19/England/QEUH-AD4C9E/2020\|EPI_ISL_634638\|20 | ···················· | ···················· | ····················· | ···················· | ···················· | ···················· |
| Q | 6541 hCoV-19/England/QEUH-AD5B54/2020\|EPI_ISL_634639\|20 | ···················· | ···················· | ····················· | ···················· | ···················· | ···················· |
| Q | 6542 hCoV-19/England/QEUH-AD578D/2020\|EPI_ISL_634640\|20 | ···················· | ···················· | ····················· | ···················· | ···················· | ···················· |
| Q | 6543 hCoV-19/England/QEUH-AD5A84/2020\|EPI_ISL_634641\|20 | ···················· | ···················· | ····················· | ···················· | ···················· | ···················· |
| Q | 6544 hCoV-19/England/QEUH-AD35D0/2020\|EPI_ISL_634642\|20 | ···················· | ···················· | ····················· | ···················· | ···················· | ···················· |
| Q | 6545 hCoV-19/England/QEUH-AD1F4D/2020\|EPI_ISL_634643\|20 | ···················· | ···················· | ····················· | ···················· | ···················· | ···················· |
| Q | 6546 hCoV-19/England/QEUH-AD1AA6/2020\|EPI_ISL_634644\|20 | ···················· | ···················· | ····················· | ···················· | ···················· | ···················· |
| Q | 6547 hCoV-19/England/QEUH-AD0FE4/2020\|EPI_ISL_634645\|20 | ···················· | ···················· | ····················· | ···················· | ···················· | ···················· |
| Q | 6548 hCoV-19/England/QEUH-AD587B/2020\|EPI_ISL_634646\|20 | ···················· | ···················· | ····················· | ···················· | ···················· | ···················· |
| Q | 6549 hCoV-19/England/QEUH-AD197C/2020\|EPI_ISL_634647\|20 | ···················· | ···················· | ····················· | ···················· | ···················· | ···················· |
| Q | 6550 hCoV-19/England/QEUH-AD4C16/2020\|EPI_ISL_634648\|20 | ···················· | ···················· | ····················· | ···················· | ···················· | ···················· |
| Q | 6551 hCoV-19/England/QEUH-AD540E/2020\|EPI_ISL_634649\|20 | ···················· | ···················· | ····················· | ···················· | ···················· | ···················· |
| Q | 6552 hCoV-19/England/QEUH-AD5A66/2020\|EPI_ISL_634650\|20 | ···················· | ···················· | ····················· | ···················· | ···················· | ···················· |
| Q | 6553 hCoV-19/England/QEUH-AD16DF/2020\|EPI_ISL_634651\|20 | ···················· | ···················· | ····················· | ···················· | ···················· | ···················· |
| Q | 6554 hCoV-19/England/QEUH-AD4C8F/2020\|EPI_ISL_634652\|20 | ···················· | ···················· | ····················· | ···················· | ···················· | ···················· |
| Q | 6555 hCoV-19/England/QEUH-AD1E7D/2020\|EPI_ISL_634653\|20 | ···················· | ···················· | ····················· | ···················· | ···················· | ···················· |
| Q | 6556 hCoV-19/England/QEUH-AD3664/2020\|EPI_ISL_634654\|20 | ···················· | ···················· | ····················· | ···················· | ···················· | ···················· |
| Q | 6557 hCoV-19/England/QEUH-AD199A/2020\|EPI_ISL_634655\|20 | ···················· | ···················· | ····················· | ···················· | ···················· | ···················· |
| Q | 6558 hCoV-19/England/QEUH-AD1F6B/2020\|EPI_ISL_634656\|20 | ···················· | ···················· | ····················· | ···················· | ···················· | ···················· |
| Q | 6559 hCoV-19/England/QEUH-AD41CC/2020\|EPI_ISL_634657\|20 | ···················· | ···················· | ····················· | ···················· | ···················· | ···················· |
| Q | 6560 hCoV-19/England/QEUH-AD4C52/2020\|EPI_ISL_634658\|20 | ···················· | ···················· | ····················· | ···················· | ···················· | ···················· |
| Q | 6561 hCoV-19/England/QEUH-AD4C25/2020\|EPI_ISL_634659\|20 | ···················· | ···················· | ····················· | ···················· | ···················· | ···················· |
| Q | 6562 hCoV-19/England/QEUH-AD4C07/2020\|EPI_ISL_634660\|20 | ···················· | ···················· | ····················· | ···················· | ···················· | ···················· |
| Q | 6563 hCoV-19/England/QEUH-AD1A00/2020\|EPI_ISL_634661\|20 | ···················· | ···················· | ····················· | ···················· | ······T············· | ···················· |
| Q | 6564 hCoV-19/England/QEUH-AD5C9D/2020\|EPI_ISL_634662\|20 | ···················· | ···················· | ····················· | ···················· | ···················· | ···················· |
| Q | 6565 hCoV-19/England/QEUH-AD3F1E/2020\|EPI_ISL_634663\|20 | ···················· | ···················· | ····················· | ···················· | ···················· | ···················· |
| Q | 6566 hCoV-19/England/QEUH-AD19D6/2020\|EPI_ISL_634664\|20 | ···················· | ···················· | ····················· | ···················· | ···················· | ···················· |
| Q | 6567 hCoV-19/England/QEUH-AD1736/2020\|EPI_ISL_634666\|20 | ···················· | ···················· | ····················· | ···················· | ···················· | ···················· |
| Q | 6568 hCoV-19/England/QEUH-AD5A93/2020\|EPI_ISL_634667\|20 | ···················· | ···················· | ····················· | ···················· | ···················· | ···················· |
| Q | 6569 hCoV-19/England/QEUH-AD35B2/2020\|EPI_ISL_634668\|20 | ···················· | ···················· | ····················· | ···················· | ···················· | ···················· |
| Q | 6570 hCoV-19/England/QEUH-AD5B45/2020\|EPI_ISL_634669\|20 | ···················· | ···················· | ····················· | ···················· | ···················· | ···················· |
| Q | 6571 hCoV-19/England/QEUH-AD42D8/2020\|EPI_ISL_634670\|20 | ···················· | ···················· | ····················· | ···················· | ···················· | ···················· |
| Q | 6572 hCoV-19/England/QEUH-AD5B81/2020\|EPI_ISL_634671\|20 | ···················· | ···················· | ····················· | ···················· | ···················· | ···················· |
| Q | 6573 hCoV-19/England/QEUH-AD1077/2020\|EPI_ISL_634672\|20 | ···················· | ···················· | ····················· | ···················· | ···················· | ···················· |
| Q | 6574 hCoV-19/England/QEUH-AD43B7/2020\|EPI_ISL_634673\|20 | ···················· | ···················· | ····················· | ···················· | ···················· | ···················· |
| Q | 6575 hCoV-19/England/QEUH-AD5B63/2020\|EPI_ISL_634674\|20 | ···················· | ·A·················· | ····················· | ···················· | ···················· | ···················· |
| Q | 6576 hCoV-19/England/QEUH-AD0F99/2020\|EPI_ISL_634675\|20 | ···················· | ···················· | ····················· | ···················· | ···················· | ···A················ |
| Q | 6577 hCoV-19/England/QEUH-AD46AF/2020\|EPI_ISL_634676\|20 | ···················· | ···················· | ····················· | ···················· | ···················· | ···················· |
| Q | 6578 hCoV-19/England/QEUH-AD1912/2020\|EPI_ISL_634677\|20 | ···················· | ···················· | ····················· | ···················· | ···················· | ···················· |
| Q | 6579 hCoV-19/England/QEUH-AD5C33/2020\|EPI_ISL_634679\|20 | ···················· | ···················· | ····················· | ···················· | ···················· | ···················· |
| Q | 6580 hCoV-19/England/QEUH-AD4469/2020\|EPI_ISL_634680\|20 | ···················· | ···················· | ····················· | ···················· | ···················· | ···················· |
| Q | 6581 hCoV-19/England/QEUH-AD586C/2020\|EPI_ISL_634681\|20 | ···················· | ···················· | ····················· | ···················· | ···················· | ···················· |
| Q | 6582 hCoV-19/England/QEUH-AD1F3E/2020\|EPI_ISL_634682\|20 | ···················· | ···················· | ····················· | ···················· | ···················· | ···················· |
| Q | 6583 hCoV-19/England/QEUH-AD4232/2020\|EPI_ISL_634683\|20 | ···················· | ···················· | ····················· | ···················· | ···················· | ···················· |
| Q | 6584 hCoV-19/England/QEUH-AD5C60/2020\|EPI_ISL_634684\|20 | ···················· | ···················· | ····················· | ···················· | ···················· | ···················· |
| Q | 6585 hCoV-19/England/QEUH-AD5AEE/2020\|EPI_ISL_634685\|20 | ···················· | ·A·················· | ····················· | ···················· | ···················· | ···················· |
| Q | 6586 hCoV-19/England/QEUH-AD1675/2020\|EPI_ISL_634686\|20 | ···················· | ···················· | ····················· | ···················· | ···················· | ···················· |
| Q | 6587 hCoV-19/England/QEUH-AD1E8C/2020\|EPI_ISL_634687\|20 | ···················· | ···················· | ····················· | ···················· | ···················· | ···················· |
| Q | 6588 hCoV-19/England/QEUH-AD4038/2020\|EPI_ISL_634688\|20 | ···················· | ···················· | ····················· | ···················· | ···················· | ···················· |
| Q | 6589 hCoV-19/England/QEUH-AD3F0F/2020\|EPI_ISL_634689\|20 | ···················· | ·A·······A·········· | ····················· | ···················· | ···················· | ···················· |
| Q | 6590 hCoV-19/England/QEUH-AD5750/2020\|EPI_ISL_634690\|20 | ···················· | ·A·················· | ····················· | ···················· | ···················· | ···A················ |
| Q | 6591 hCoV-19/England/QEUH-AD5741/2020\|EPI_ISL_634691\|20 | ···················· | ···················· | ····················· | ···················· | ···················· | ···················· |
| Q | 6592 hCoV-19/England/QEUH-AD3F78/2020\|EPI_ISL_634692\|20 | ···················· | ···················· | ····················· | ···················· | ···················· | ···················· |
| Q | 6593 hCoV-19/England/QEUH-AD5A48/2020\|EPI_ISL_634693\|20 | ···················· | ···················· | ····················· | ···················· | ···················· | ···················· |
| Q | 6594 hCoV-19/England/QEUH-AD1930/2020\|EPI_ISL_634694\|20 | ···················· | ···················· | ····················· | ···················· | ···················· | ···················· |
| Q | 6595 hCoV-19/England/QEUH-AD3FE1/2020\|EPI_ISL_634695\|20 | ···················· | ···················· | ····················· | ···················· | ···················· | ···················· |
| Q | 6596 hCoV-19/England/QEUH-AD46DC/2020\|EPI_ISL_634696\|20 | ···················· | ···················· | ····················· | ···················· | ···················· | ···················· |
| Q | 6597 hCoV-19/England/QEUH-AD442D/2020\|EPI_ISL_634697\|20 | ···················· | ···················· | ····················· | ···················· | ···················· | ···················· |
| Q | 6598 hCoV-19/England/QEUH-AD437B/2020\|EPI_ISL_634698\|20 | ···················· | ···················· | ····················· | ···················· | ···················· | ···················· |
| Q | 6599 hCoV-19/England/QEUH-AD1F98/2020\|EPI_ISL_634699\|20 | ···················· | ···················· | ····················· | ···················· | ···················· | ···················· |
| Q | 6600 hCoV-19/South Africa/KRISP-K004029/2020\|EPI_ISL_63 | ···················· | ···················· | ····················· | ···················· | ···················· | ···················· |

| | | | | | | | |
|---|---|---|---|---|---|---|---|
| Q | 6601 hCoV-19/South Africa/KRISP-K004030/2020\|EPI_ISL_63 | ···················· | ···················· | ····················· | ···················· | ···················· | ···················· |
| Q | 6602 hCoV-19/South Africa/KRISP-K004031/2020\|EPI_ISL_63 | ···················· | ···················· | ····················· | ···················· | ···················· | ···················· |
| Q | 6603 hCoV-19/South Africa/KRISP-K004040/2020\|EPI_ISL_63 | ···················· | ·A·················· | ····················· | ···················· | ···················· | ···················· |
| Q | 6604 hCoV-19/South Africa/KRISP-K004041/2020\|EPI_ISL_63 | ···················· | ···················· | ····················· | ···················· | ···················· | ···················· |
| Q | 6605 hCoV-19/South Africa/KRISP-K004045/2020\|EPI_ISL_63 | ···················· | ···················· | ····················· | ···················· | ···················· | ···················· |
| Q | 6606 hCoV-19/South Africa/KRISP-K004046/2020\|EPI_ISL_63 | ···················· | ···················· | ····················· | ···················· | ···················· | ···················· |
| Q | 6607 hCoV-19/South Africa/KRISP-K004047/2020\|EPI_ISL_63 | ···················· | ···················· | ····················· | ···················· | ···················· | ···················· |
| Q | 6608 hCoV-19/South Africa/KRISP-K004048/2020\|EPI_ISL_63 | ···················· | ···················· | ····················· | ···················· | ···················· | ···················· |
| Q | 6609 hCoV-19/South Africa/KRISP-K004049/2020\|EPI_ISL_63 | ···················· | ·A·················· | ····················· | ···················· | ···················· | ···················· |
| Q | 6610 hCoV-19/South Africa/KRISP-K004050/2020\|EPI_ISL_63 | ···················· | ·A·················· | ····················· | ···················· | ···················· | ···················· |
| Q | 6611 hCoV-19/USA/CA-ALSR-3175/2020\|EPI_ISL_635901\|2020- | ········T··········· | ···················· | ····················· | ···················· | ···················· | ···················· |
| Q | 6612 hCoV-19/USA/CA-ALSR-3178/2020\|EPI_ISL_635902\|2020- | ···················· | ·A·················· | ····················· | ···················· | ···················· | ···················· |
| Q | 6613 hCoV-19/USA/CA-ALSR-3181/2020\|EPI_ISL_635903\|2020- | ···················· | ·A·················· | ····················· | ···················· | ···················· | ···················· |
| Q | 6614 hCoV-19/USA/CA-ALSR-3186/2020\|EPI_ISL_635904\|2020- | ···················· | ···················· | ····················· | ···················· | ···················· | ···················· |
| Q | 6615 hCoV-19/USA/CA-ALSR-3197/2020\|EPI_ISL_635909\|2020- | ···················· | ···················· | ····················· | ···················· | ···················· | ···················· |
| Q | 6616 hCoV-19/USA/CA-ALSR-3331/2020\|EPI_ISL_635967\|2020- | ···················· | ·A·················· | ····················· | ···················· | ···················· | ···················· |
| Q | 6617 hCoV-19/USA/CA-ALSR-3332/2020\|EPI_ISL_635968\|2020- | ···················· | ·A·················· | ····················· | ···················· | ···················· | ···················· |
| Q | 6618 hCoV-19/USA/CA-ALSR-3337/2020\|EPI_ISL_635969\|2020- | ···················· | ·A·················· | ····················· | ···················· | ···················· | ···················· |
| Q | 6619 hCoV-19/USA/CA-ALSR-3341/2020\|EPI_ISL_635972\|2020- | ···················· | ·A·················· | ····················· | ···················· | ···················· | ···················· |
| Q | 6620 hCoV-19/USA/CA-ALSR-3345/2020\|EPI_ISL_635973\|2020- | ···················· | ·A·················· | ····················· | ···················· | ···················· | ···················· |
| Q | 6621 hCoV-19/USA/CA-ALSR-3349/2020\|EPI_ISL_635974\|2020- | ···················· | ·A·················· | ····················· | ···················· | ···················· | ···················· |
| Q | 6622 hCoV-19/USA/CA-ALSR-3351/2020\|EPI_ISL_635975\|2020- | ···················· | ·A·················· | ····················· | ···················· | ···················· | ···················· |
| Q | 6623 hCoV-19/USA/CA-ALSR-3352/2020\|EPI_ISL_635976\|2020- | ···················· | ·A·················· | ····················· | ···················· | ···················· | ···················· |
| Q | 6624 hCoV-19/USA/CA-ALSR-3354/2020\|EPI_ISL_635977\|2020- | ···················· | ·A·················· | ····················· | ···················· | ···················· | ···················· |
| Q | 6625 hCoV-19/USA/CA-ALSR-3358/2020\|EPI_ISL_635980\|2020- | ···················· | ·A·················· | ····················· | ···················· | ···················· | ···················· |
| Q | 6626 hCoV-19/USA/CA-ALSR-3360/2020\|EPI_ISL_635981\|2020- | ···················· | ·A·················· | ····················· | ···················· | ···················· | ···················· |
| Q | 6627 hCoV-19/USA/CA-ALSR-3362/2020\|EPI_ISL_635982\|2020- | ···················· | ·A·················· | ····················· | ···················· | ···················· | ···················· |
| Q | 6628 hCoV-19/USA/CA-ALSR-3366/2020\|EPI_ISL_635983\|2020- | ···················· | ·A·················· | ····················· | ···················· | ···················· | ···················· |
| Q | 6629 hCoV-19/USA/CA-ALSR-3369/2020\|EPI_ISL_635984\|2020- | ···················· | ···················· | ····················· | ···················· | ···················· | ···················· |
| Q | 6630 hCoV-19/USA/CA-ALSR-3370/2020\|EPI_ISL_635985\|2020- | ···················· | ·A·················· | ····················· | ···················· | ···················· | ···················· |
| Q | 6631 hCoV-19/USA/CA-ALSR-3373/2020\|EPI_ISL_635986\|2020- | ···················· | ·A·················· | ····················· | ···················· | ···················· | ···················· |
| Q | 6632 hCoV-19/USA/CA-ALSR-3374/2020\|EPI_ISL_635987\|2020- | ···················· | ·A·················· | ····················· | ···················· | ···················· | ···················· |
| Q | 6633 hCoV-19/USA/CA-ALSR-3375/2020\|EPI_ISL_635988\|2020- | ···················· | ·A·················· | ····················· | ···················· | ···················· | ···················· |
| Q | 6634 hCoV-19/USA/CA-ALSR-3376/2020\|EPI_ISL_635989\|2020- | ···················· | ·A·················· | ····················· | ···················· | ···················· | ···················· |
| Q | 6635 hCoV-19/USA/CA-ALSR-3377/2020\|EPI_ISL_635990\|2020- | ···················· | ···················· | ····················· | ···················· | ···················· | ···················· |
| Q | 6636 hCoV-19/USA/CA-ALSR-3380/2020\|EPI_ISL_635991\|2020- | ···················· | ···················· | ····················· | ···················· | ···················· | ···················· |
| Q | 6637 hCoV-19/USA/CA-ALSR-3381/2020\|EPI_ISL_635992\|2020- | ···················· | ·A·················· | ····················· | ···················· | ···················· | ···················· |
| Q | 6638 hCoV-19/USA/CA-ALSR-3382/2020\|EPI_ISL_635993\|2020- | ···················· | ·A·················· | ····················· | ···················· | ···················· | ···················· |
| Q | 6639 hCoV-19/USA/CA-ALSR-3384/2020\|EPI_ISL_635994\|2020- | ···················· | ·A·················· | ····················· | ···················· | ···················· | ···················· |
| Q | 6640 hCoV-19/USA/CA-ALSR-3388/2020\|EPI_ISL_635996\|2020- | ···················· | ·A·················· | ····················· | ···················· | ···················· | ···················· |
| Q | 6641 hCoV-19/USA/CA-ALSR-3389/2020\|EPI_ISL_635997\|2020- | ········T··········· | ·A·················· | ····················· | ···················· | ···················· | ···················· |
| Q | 6642 hCoV-19/USA/CA-ALSR-3391/2020\|EPI_ISL_635998\|2020- | ···················· | ·A·················· | ····················· | ···················· | ···················· | ···················· |
| Q | 6643 hCoV-19/USA/CA-ALSR-3392/2020\|EPI_ISL_635999\|2020- | ···················· | ···················· | ····················· | ···················· | ···················· | ···················· |
| Q | 6644 hCoV-19/Jordan/AM-ALSR-3981/2020\|EPI_ISL_636344\|20 | ···················· | ·A·················· | ····················· | ···················· | ···················· | ···A················ |
| Q | 6645 hCoV-19/Jordan/AM-ALSR-3982/2020\|EPI_ISL_636345\|20 | ···················· | ···················· | ····················· | ···················· | ···················· | ···················· |
| Q | 6646 hCoV-19/Jordan/AM-ALSR-3983/2020\|EPI_ISL_636346\|20 | ···················· | ···················· | ····················· | ···················· | ···················· | ···················· |
| Q | 6647 hCoV-19/Jordan/AM-ALSR-3987/2020\|EPI_ISL_636350\|20 | ···················· | ···················· | ····················· | ···················· | ···················· | ···················· |
| Q | 6648 hCoV-19/Jordan/AM-ALSR-3988/2020\|EPI_ISL_636351\|20 | ···················· | ···················· | ····················· | ···················· | ···················· | ···················· |
| Q | 6649 hCoV-19/Jordan/AM-ALSR-3989/2020\|EPI_ISL_636352\|20 | ···················· | ···················· | ····················· | ···················· | ···················· | ···················· |
| Q | 6650 hCoV-19/Jordan/AM-ALSR-3990/2020\|EPI_ISL_636353\|20 | ···················· | ···················· | ····················· | ···················· | ···················· | ···················· |
| Q | 6651 hCoV-19/Jordan/AM-ALSR-3991/2020\|EPI_ISL_636354\|20 | ···················· | ···················· | ····················· | ···················· | ···················· | ···················· |
| Q | 6652 hCoV-19/Jordan/AM-ALSR-3992/2020\|EPI_ISL_636355\|20 | ···················· | ···················· | ····················· | ···················· | ···················· | ···················· |
| Q | 6653 hCoV-19/Jordan/AM-ALSR-3993/2020\|EPI_ISL_636356\|20 | ···················· | ·A·················· | ····················· | ···················· | ···················· | ···A················ |
| Q | 6654 hCoV-19/Jordan/AM-ALSR-3994/2020\|EPI_ISL_636357\|20 | ···················· | ···················· | ····················· | ···················· | ···················· | ···················· |
| Q | 6655 hCoV-19/Jordan/AM-ALSR-3995/2020\|EPI_ISL_636358\|20 | ···················· | ···················· | ····················· | ···················· | ···················· | ···················· |
| Q | 6656 hCoV-19/Jordan/AM-ALSR-3996/2020\|EPI_ISL_636359\|20 | ···················· | ···················· | ····················· | ···················· | ···················· | ···················· |
| Q | 6657 hCoV-19/England/MILK-ACEBD6/2020\|EPI_ISL_629140\|20 | ···················· | ····G··············· | ····················· | ···················· | ···················· | ···················· |
| Q | 6658 hCoV-19/England/MILK-AC834D/2020\|EPI_ISL_629141\|20 | ···················· | ···················· | ····················· | ···················· | ···················· | ···················· |
| Q | 6659 hCoV-19/England/MILK-ACBF91/2020\|EPI_ISL_629200\|20 | ···················· | ···················· | ····················· | ···················· | ···················· | ···················· |
| Q | 6660 hCoV-19/England/MILK-ACC1D6/2020\|EPI_ISL_629201\|20 | ···················· | ·A·················· | ····················· | ···················· | ···················· | ···A················ |
| Q | 6661 hCoV-19/England/MILK-ACE981/2020\|EPI_ISL_629202\|20 | ···················· | ·A·················· | ····················· | ···················· | ···················· | ···A················ |
| Q | 6662 hCoV-19/England/MILK-ACBE85/2020\|EPI_ISL_629203\|20 | ···················· | ···················· | ····················· | ···················· | ···················· | ···················· |
| Q | 6663 hCoV-19/England/MILK-AC8635/2020\|EPI_ISL_629204\|20 | ···················· | ···················· | ····················· | ···················· | ···················· | ···················· |
| Q | 6664 hCoV-19/England/MILK-ACC23D/2020\|EPI_ISL_629205\|20 | ···················· | ···················· | ····················· | ···················· | ···················· | ···················· |
| Q | 6665 hCoV-19/England/MILK-ACE392/2020\|EPI_ISL_629206\|20 | ···················· | ···················· | ····················· | ···················· | ···················· | ···················· |
| Q | 6666 hCoV-19/England/MILK-AC82D7/2020\|EPI_ISL_629207\|20 | ···················· | ···················· | ····················· | ···················· | ···················· | ···················· |
| Q | 6667 hCoV-19/England/MILK-ACC024/2020\|EPI_ISL_629208\|20 | ···················· | ···················· | ····················· | ···················· | ······T············· | ···················· |
| Q | 6668 hCoV-19/England/MILK-ACE1C5/2020\|EPI_ISL_630127\|20 | ···················· | ···················· | ····················· | ···················· | ···················· | ···················· |
| Q | 6669 hCoV-19/England/MILK-ACDBB9/2020\|EPI_ISL_630130\|20 | ···················· | ·A·················· | ····················· | ···················· | ···················· | ···A················ |
| Q | 6670 hCoV-19/England/MILK-ACDB6E/2020\|EPI_ISL_630131\|20 | ···················· | ···················· | ····················· | ···················· | ···················· | ···················· |
| Q | 6671 hCoV-19/England/MILK-ACE12F/2020\|EPI_ISL_630132\|20 | ···················· | ···················· | ····················· | ···················· | ···················· | ···················· |
| Q | 6672 hCoV-19/England/MILK-ACD08D/2020\|EPI_ISL_630133\|20 | ···················· | ···················· | ····················· | ···················· | ···················· | ···················· |
| Q | 6673 hCoV-19/England/MILK-ACDC5C/2020\|EPI_ISL_630134\|20 | ···················· | ···················· | ····················· | ···················· | ···················· | ···················· |
| Q | 6674 hCoV-19/England/MILK-ACDBD7/2020\|EPI_ISL_630135\|20 | ···················· | ···················· | ····················· | ···················· | ···················· | ···················· |
| Q | 6675 hCoV-19/England/MILK-ACDB13/2020\|EPI_ISL_630136\|20 | ···················· | ···················· | ····················· | ···················· | ···················· | ···················· |
| Q | 6676 hCoV-19/USA/WI-WSLH-200701/2020\|EPI_ISL_631458\|202 | ···················· | ···················· | ····················· | ···················· | ···················· | ···················· |
| Q | 6679 hCoV-19/USA/WI-WSLH-200705/2020\|EPI_ISL_631462\|202 | ···················· | ···················· | ····················· | ···················· | ···················· | ···················· |
| Q | 6680 hCoV-19/USA/WI-WSLH-200706/2020\|EPI_ISL_631463\|202 | ···················· | ···················· | ····················· | ···················· | ···················· | ···················· |
| Q | 6681 hCoV-19/USA/WI-WSLH-200707/2020\|EPI_ISL_631464\|202 | ···················· | ···················· | ····················· | ···················· | ···················· | ···················· |
| Q | 6682 hCoV-19/USA/WI-WSLH-200708/2020\|EPI_ISL_631465\|202 | ···················· | ···················· | ····················· | ···················· | ···················· | ···················· |

| | | | | | | | |
|---|---|---|---|---|---|---|---|
| Q | 6687 hCoV-19/USA/NY-NYCPHL-000001/2020\|EPI_ISL_631502\|2 | ···················· | ···················· | ···················· | ···················· | ···················· | ···················· |
| Q | 6688 hCoV-19/USA/NY-NYCPHL-000003/2020\|EPI_ISL_631504\|2 | ···················· | ···················· | ···················· | ···················· | ···················· | ···················· |
| Q | 6689 hCoV-19/USA/NY-NYCPHL-000005/2020\|EPI_ISL_631505\|2 | ···················· | ···················· | ···················· | ···················· | ···················· | ···················· |
| Q | 6690 hCoV-19/USA/NY-NYCPHL-000009/2020\|EPI_ISL_631508\|2 | ···················· | ···················· | ···················· | ···················· | ···················· | ···················· |
| Q | 6691 hCoV-19/USA/NY-NYCPHL-000010/2020\|EPI_ISL_631509\|2 | ···················· | ···················· | ···················· | ···················· | ···················· | ···················· |
| Q | 6692 hCoV-19/USA/NY-NYCPHL-000012/2020\|EPI_ISL_631510\|2 | ···················· | ···················· | ···················· | ···················· | ···················· | ···················· |
| Q | 6693 hCoV-19/USA/NY-NYCPHL-000013/2020\|EPI_ISL_631511\|2 | ···················· | ···················· | ···················· | ···················· | ···················· | ···················· |
| Q | 6694 hCoV-19/USA/NY-NYCPHL-000014/2020\|EPI_ISL_631512\|2 | ···················· | ···················· | ···················· | ···················· | ···················· | ···················· |
| Q | 6695 hCoV-19/USA/NY-NYCPHL-000015/2020\|EPI_ISL_631513\|2 | ···················· | ···················· | ···················· | ···················· | ···················· | ···················· |
| Q | 6696 hCoV-19/USA/NY-NYCPHL-000016/2020\|EPI_ISL_631514\|2 | ···················· | ···················· | ···················· | ···················· | ···················· | ···················· |
| Q | 6697 hCoV-19/USA/NY-NYCPHL-000017/2020\|EPI_ISL_631515\|2 | ···················· | ···················· | ···················· | ···················· | ···················· | ···················· |
| Q | 6698 hCoV-19/USA/NY-NYCPHL-000019/2020\|EPI_ISL_631516\|2 | ···················· | ···················· | ···················· | ···················· | ···················· | ···················· |
| Q | 6699 hCoV-19/USA/NY-NYCPHL-000021/2020\|EPI_ISL_631517\|2 | ···················· | ···················· | ···················· | ···················· | ···················· | ···················· |
| Q | 6700 hCoV-19/USA/NY-NYCPHL-000022/2020\|EPI_ISL_631518\|2 | ···················· | ···················· | ···················· | ···················· | ···················· | ···················· |
| Q | 6701 hCoV-19/USA/NY-NYCPHL-000023/2020\|EPI_ISL_631519\|2 | ···················· | ···················· | ···················· | ···················· | ···················· | ···················· |
| Q | 6702 hCoV-19/USA/NY-NYCPHL-000024/2020\|EPI_ISL_631520\|2 | ···················· | ···················· | ···················· | ···················· | ···················· | ···················· |
| Q | 6703 hCoV-19/USA/NY-NYCPHL-000026/2020\|EPI_ISL_631522\|2 | ···················· | ···················· | ···················· | ···················· | ···················· | ···················· |
| Q | 6704 hCoV-19/Netherlands/ZH-EMC-900/2020\|EPI_ISL_632790 | ···················· | ···················· | ···················· | ···················· | ···················· | ···················· |
| Q | 6705 hCoV-19/Netherlands/ZH-EMC-901/2020\|EPI_ISL_632791 | ···················· | ···················· | ···················· | ···················· | ···················· | ···················· |
| Q | 6706 hCoV-19/Netherlands/ZH-EMC-902/2020\|EPI_ISL_632792 | ···················· | ···················· | ···T················ | ···················· | ···················· | ···················· |
| Q | 6707 hCoV-19/Netherlands/ZH-EMC-903/2020\|EPI_ISL_632793 | ···················· | ···················· | ···T················ | ···················· | ···················· | ···················· |
| Q | 6708 hCoV-19/Netherlands/ZH-EMC-904/2020\|EPI_ISL_632794 | ···················· | ···················· | ···T················ | ···················· | ···················· | ···················· |
| Q | 6709 hCoV-19/Netherlands/ZH-EMC-905/2020\|EPI_ISL_632795 | ···················· | ···················· | ···T················ | ···················· | ···················· | ···················· |
| Q | 6710 hCoV-19/England/204462558/2020\|EPI_ISL_632796\|2020 | ···················· | ···················· | ···················· | ···················· | ···················· | ···················· |
| Q | 6711 hCoV-19/Gibraltar/204462899/2020\|EPI_ISL_632797\|20 | ···················· | ···················· | ···················· | ···················· | ···················· | ···················· |
| Q | 6712 hCoV-19/Gibraltar/204462900/2020\|EPI_ISL_632798\|20 | ···················· | ···················· | ···················· | ···················· | ···················· | ···················· |
| Q | 6713 hCoV-19/Gibraltar/204462902/2020\|EPI_ISL_632800\|20 | ···················· | ···················· | ···················· | ···················· | ···················· | ···················· |
| Q | 6714 hCoV-19/Gibraltar/204462903/2020\|EPI_ISL_632801\|20 | ···················· | ···················· | ···················· | ···················· | ···················· | ···················· |
| Q | 6715 hCoV-19/Gibraltar/204462904/2020\|EPI_ISL_632802\|20 | ···················· | ···················· | ···················· | ···················· | ···················· | ···················· |
| Q | 6716 hCoV-19/Gibraltar/204462905/2020\|EPI_ISL_632803\|20 | ···················· | ···················· | ···················· | ···················· | ···················· | ···················· |
| Q | 6717 hCoV-19/Gibraltar/204462906/2020\|EPI_ISL_632804\|20 | ···················· | ···················· | ···················· | ···················· | ···················· | ···················· |
| Q | 6718 hCoV-19/Gibraltar/204462907/2020\|EPI_ISL_632805\|20 | ···················· | ···················· | ···················· | ···················· | ···················· | ···················· |
| Q | 6719 hCoV-19/Gibraltar/204462908/2020\|EPI_ISL_632806\|20 | ···················· | ···················· | ···················· | ···················· | ···················· | ···················· |
| Q | 6720 hCoV-19/Gibraltar/204462909/2020\|EPI_ISL_632807\|20 | ···················· | ···················· | ···················· | ···················· | ···················· | ···················· |
| Q | 6721 hCoV-19/Gibraltar/204462910/2020\|EPI_ISL_632808\|20 | ···················· | ···················· | ···················· | ···················· | ···················· | ···················· |
| Q | 6722 hCoV-19/Gibraltar/204462911/2020\|EPI_ISL_632809\|20 | ···················· | ···················· | ···················· | ···················· | ···················· | ···················· |
| Q | 6723 hCoV-19/Gibraltar/204462912/2020\|EPI_ISL_632810\|20 | ···················· | ···················· | ···················· | ···················· | ···················· | ···················· |
| Q | 6724 hCoV-19/Gibraltar/204462913/2020\|EPI_ISL_632811\|20 | ···················· | ···················· | ···················· | ···················· | ···················· | ···················· |
| Q | 6725 hCoV-19/Gibraltar/204462914/2020\|EPI_ISL_632812\|20 | ···················· | ···················· | ···················· | ···················· | ···················· | ···················· |
| Q | 6726 hCoV-19/Gibraltar/204462915/2020\|EPI_ISL_632813\|20 | ···················· | ···················· | ···················· | ···················· | ···················· | ···················· |
| Q | 6727 hCoV-19/Gibraltar/204462917/2020\|EPI_ISL_632815\|20 | ···················· | ···················· | ···················· | ···················· | ···················· | ···················· |
| Q | 6728 hCoV-19/Gibraltar/204462918/2020\|EPI_ISL_632816\|20 | ···················· | ···················· | ···················· | ···················· | ······T············· | ···················· |
| Q | 6729 hCoV-19/Gibraltar/204462920/2020\|EPI_ISL_632817\|20 | ···················· | ···················· | ···················· | ···················· | ···················· | ···················· |
| Q | 6730 hCoV-19/Gibraltar/204462921/2020\|EPI_ISL_632818\|20 | ···················· | ···················· | ···················· | ···················· | ···················· | ···················· |
| Q | 6731 hCoV-19/Gibraltar/204462922/2020\|EPI_ISL_632819\|20 | ···················· | ···················· | ···················· | ···················· | ···················· | ···················· |
| Q | 6732 hCoV-19/Gibraltar/204462923/2020\|EPI_ISL_632820\|20 | ···················· | ···················· | ···················· | ···················· | ···················· | ···················· |
| Q | 6733 hCoV-19/Gibraltar/204462924/2020\|EPI_ISL_632821\|20 | ···················· | ···················· | ···················· | ···················· | ······T············· | ···················· |
| Q | 6734 hCoV-19/Gibraltar/204462925/2020\|EPI_ISL_632822\|20 | ···················· | ···················· | ···················· | ···················· | ···················· | ···················· |
| Q | 6735 hCoV-19/USA/NM-NMDOH-2020320996/2020\|EPI_ISL_63282 | ···················· | ···················· | ···················· | ···················· | ···················· | ···················· |
| Q | 6736 hCoV-19/USA/NM-NMDOH-2020324072/2020\|EPI_ISL_63282 | ···················· | ···················· | ···················· | ···················· | ···················· | ···················· |
| Q | 6737 hCoV-19/USA/NM-NMDOH-2020324112/2020\|EPI_ISL_63282 | ···················· | ···················· | ···················· | ···················· | ···················· | ···················· |
| Q | 6738 hCoV-19/USA/NM-NMDOH-2020324210/2020\|EPI_ISL_63282 | ···················· | ···················· | ···················· | ···················· | ···················· | ···················· |
| Q | 6739 hCoV-19/USA/NM-NMDOH-2020324222/2020\|EPI_ISL_63282 | ···················· | ···················· | ···················· | ···················· | ···················· | ···················· |
| Q | 6740 hCoV-19/USA/NM-NMDOH-2020324268/2020\|EPI_ISL_63282 | ···················· | ···················· | ···················· | ···················· | ···················· | ···················· |
| Q | 6741 hCoV-19/USA/NM-NMDOH-2020324418/2020\|EPI_ISL_63283 | ···················· | ···················· | ···················· | ···················· | ···················· | ···················· |
| Q | 6742 hCoV-19/USA/NM-NMDOH-2020324473/2020\|EPI_ISL_63283 | ···················· | ···················· | ···················· | ···················· | ···················· | ···················· |
| Q | 6743 hCoV-19/USA/NM-NMDOH-2020324645/2020\|EPI_ISL_63283 | ···················· | ···················· | ···················· | ···················· | ···················· | ···················· |
| Q | 6744 hCoV-19/USA/NM-NMDOH-2020324951/2020\|EPI_ISL_63283 | ···················· | ···················· | ···················· | ···················· | ···················· | ···················· |
| Q | 6745 hCoV-19/USA/NM-NMDOH-2020324977/2020\|EPI_ISL_63283 | ···················· | ···················· | ···················· | ···················· | ···················· | ···················· |
| Q | 6746 hCoV-19/USA/NM-NMDOH-2020325112/2020\|EPI_ISL_63283 | ···················· | ···················· | ···················· | ···················· | ···················· | ···················· |
| Q | 6747 hCoV-19/USA/NM-NMDOH-2020326528/2020\|EPI_ISL_63284 | ···················· | ···················· | ···················· | ···················· | ···················· | ···················· |
| Q | 6748 hCoV-19/USA/NM-NMDOH-2020326664/2020\|EPI_ISL_63284 | ···················· | ···················· | ···················· | ···················· | ···················· | ···················· |
| Q | 6749 hCoV-19/USA/NM-NMDOH-2020326676/2020\|EPI_ISL_63284 | ···················· | ···················· | ···················· | ···················· | ···················· | ···················· |
| Q | 6750 hCoV-19/USA/NM-NMDOH-2020326697/2020\|EPI_ISL_63284 | ···················· | ···················· | ···················· | ···················· | ···················· | ···················· |
| Q | 6751 hCoV-19/USA/ID-UNM-IBL032/2020\|EPI_ISL_632847\|2020 | ···················· | ···················· | ···················· | ···················· | ···················· | ···················· |
| Q | 6752 hCoV-19/USA/ID-UNM-IBL031/2020\|EPI_ISL_632848\|2020 | ···················· | ···················· | ···················· | ···················· | ···················· | ···················· |
| Q | 6753 hCoV-19/USA/ID-UNM-IBL055/2020\|EPI_ISL_632849\|2020 | ···················· | ···················· | ···················· | ···················· | ···················· | ···················· |
| Q | 6754 hCoV-19/USA/ID-UNM-IBL030/2020\|EPI_ISL_632850\|2020 | ···················· | ···················· | ···················· | ···················· | ···················· | ···················· |
| Q | 6755 hCoV-19/USA/ID-UNM-IBL312/2020\|EPI_ISL_632851\|2020 | ···················· | ···················· | ···················· | ···················· | ···················· | ···················· |
| Q | 6756 hCoV-19/USA/ID-UNM-IBL033/2020\|EPI_ISL_632852\|2020 | ···················· | ···················· | ···················· | ···················· | ···················· | ···················· |
| Q | 6757 hCoV-19/USA/ID-UNM-IBL053/2020\|EPI_ISL_632853\|2020 | ···················· | ···················· | ···················· | ···················· | ···················· | ···················· |
| Q | 6758 hCoV-19/South Africa/KRISP-K004015/2020\|EPI_ISL_63 | ···················· | ···················· | ···················· | ···················· | ···················· | ···················· |
| Q | 6759 hCoV-19/South Africa/KRISP-K004017/2020\|EPI_ISL_63 | ···················· | ···················· | ···················· | ···················· | ···················· | ···················· |
| Q | 6760 hCoV-19/South Africa/KRISP-K004018/2020\|EPI_ISL_63 | ······G············· | ···················· | ···················· | ···················· | ···················· | ···················· |
| Q | 6761 hCoV-19/South Africa/KRISP-K004020/2020\|EPI_ISL_63 | ···················· | ···················· | ···················· | ···················· | ···················· | ···················· |
| Q | 6762 hCoV-19/South Africa/KRISP-K004021/2020\|EPI_ISL_63 | ···················· | ···················· | ···················· | ···················· | ···················· | ···················· |
| Q | 6763 hCoV-19/South Africa/KRISP-K004022/2020\|EPI_ISL_63 | ···················· | ···················· | ···················· | ···················· | ···················· | ···················· |
| Q | 6764 hCoV-19/South Africa/KRISP-K004023/2020\|EPI_ISL_63 | ···················· | ···················· | ···················· | ···················· | ···················· | ···················· |
| Q | 6765 hCoV-19/South Africa/KRISP-K004024/2020\|EPI_ISL_63 | ···················· | ·A·················· | ···················· | ···················· | ···················· | ···················· |
| Q | 6766 hCoV-19/South Africa/KRISP-K004025/2020\|EPI_ISL_63 | ···················· | ···················· | ···················· | ···················· | ···················· | ···················· |

Q 6767 hCoV-19/South Africa/KRISP-K004027/2020|EPI_ISL_63 ···················· ···················· ···················· ···················· ···················· ····················
Q 6768 hCoV-19/South Africa/KRISP-K004032/2020|EPI_ISL_63 ···················· ···················· ···················· ···················· ···················· ····················
Q 6769 hCoV-19/South Africa/KRISP-K004033/2020|EPI_ISL_63 ···················· ···················· ···················· ···················· ···················· ····················
Q 6770 hCoV-19/South Africa/KRISP-K004034/2020|EPI_ISL_63 ···················· ···················· ···················· ···················· ···················· ····················
Q 6771 hCoV-19/South Africa/KRISP-K004035/2020|EPI_ISL_63 ···················· ···················· ···················· ···················· ···················· ····················
Q 6772 hCoV-19/South Africa/KRISP-K004036/2020|EPI_ISL_63 ···················· ·A·················· ···················· ···················· ···················· ····················
Q 6773 hCoV-19/South Africa/KRISP-K004037/2020|EPI_ISL_63 ···················· ···················· ···················· ···················· ···················· ····················
Q 6774 hCoV-19/South Africa/KRISP-K004038/2020|EPI_ISL_63 ···················· ···················· ···················· ···················· ···················· ····················
Q 6775 hCoV-19/South Africa/KRISP-K004051/2020|EPI_ISL_63 ···················· ···················· ···················· ···················· ···················· ····················
Q 6776 hCoV-19/South Africa/KRISP-K004052/2020|EPI_ISL_63 ···················· ···················· ···················· ···················· ···················· ····················
Q 6777 hCoV-19/South Africa/KRISP-K004053/2020|EPI_ISL_63 ···················· ·A·················· ···················· ···················· ···················· ····················
Q 6778 hCoV-19/South Africa/KRISP-K004054/2020|EPI_ISL_63 ··········T········· ···················· ···················· ···················· ···················· ····················
Q 6779 hCoV-19/South Africa/KRISP-K004055/2020|EPI_ISL_63 ···················· ·A·················· ···················· ···················· ···················· ····················
Q 6780 hCoV-19/South Africa/KRISP-K004056/2020|EPI_ISL_63 ···················· ···················· ···················· ···················· ···················· ····················
Q 6781 hCoV-19/South Africa/KRISP-K004057/2020|EPI_ISL_63 ···················· ·A·················· ···················· ···················· ···················· ····················
Q 6782 hCoV-19/South Africa/KRISP-K004058/2020|EPI_ISL_63 ···················· ·A·················· ···················· ···················· ···················· ····················
Q 6783 hCoV-19/South Africa/KRISP-K004059/2020|EPI_ISL_63 ···················· ·A·················· ···················· ···················· ···················· ····················
Q 6784 hCoV-19/South Africa/KRISP-K004060/2020|EPI_ISL_63 ···················· ···················· ···················· ···················· ···················· ····················
Q 6785 hCoV-19/South Africa/KRISP-K004061/2020|EPI_ISL_63 ···················· ·A·················· ···················· ···················· ···················· ····················
Q 6786 hCoV-19/South Africa/KRISP-K004062/2020|EPI_ISL_63 ···················· ···················· ···················· ···················· ···················· ····················
Q 6787 hCoV-19/South Africa/KRISP-K004064/2020|EPI_ISL_63 ···················· ···················· ···················· ···················· ···················· ····················
Q 6788 hCoV-19/South Africa/KRISP-K004066/2020|EPI_ISL_63 ···················· ···················· ···················· ···················· ···················· ····················
Q 6790 hCoV-19/South Africa/KRISP-K004069/2020|EPI_ISL_63 ···················· ···················· ···················· ···················· ···················· ····················
Q 6791 hCoV-19/South Africa/KRISP-K004070/2020|EPI_ISL_63 ···················· ···················· ···················· ···················· ···················· ····················
Q 6792 hCoV-19/South Africa/KRISP-K004071/2020|EPI_ISL_63 ···················· ···················· ···················· ···················· ···················· ····················
Q 6793 hCoV-19/South Africa/KRISP-K004072/2020|EPI_ISL_63 ···················· ·A·················· ···················· ···················· ···················· ····················
Q 6794 hCoV-19/South Africa/KRISP-K004074/2020|EPI_ISL_63 ···················· ·A·················· ···················· ···················· ···················· ····················
Q 6795 hCoV-19/South Africa/KRISP-K004075/2020|EPI_ISL_63 ···················· ···················· ···················· ···················· ···················· ····················
Q 6796 hCoV-19/South Africa/KRISP-K004077/2020|EPI_ISL_63 ···················· ···················· ···················· ···················· ···················· ····················
Q 6797 hCoV-19/South Africa/KRISP-K004078/2020|EPI_ISL_63 ···················· ···················· ···················· ···················· ···················· ····················
Q 6798 hCoV-19/South Africa/KRISP-K004079/2020|EPI_ISL_63 ···················· ···················· ···················· ···················· ···················· ····················
Q 6799 hCoV-19/South Africa/KRISP-K004080/2020|EPI_ISL_63 ···················· ···················· ···················· ···················· ···················· ····················
Q 6800 hCoV-19/South Africa/KRISP-K004081/2020|EPI_ISL_63 ···················· ···················· ···················· ···················· ···················· ····················
Q 6801 hCoV-19/South Africa/KRISP-K004082/2020|EPI_ISL_63 ···················· ·A·················· ···················· ···················· ···················· ····················
Q 6802 hCoV-19/Tunisia/55400/2020|EPI_ISL_635061|2020-11- ···················· ···················· ···················· ···················· ···················· ····················
Q 6803 hCoV-19/Norway/4598/2020|EPI_ISL_635063|2020-10-02 ···················· ···················· ···················· ···················· ···················· ····················
Q 6804 hCoV-19/Norway/4599/2020|EPI_ISL_635064|2020-09-30 ···················· ···················· ···················· ···················· ···················· ····················
Q 6805 hCoV-19/Norway/4617/2020|EPI_ISL_635065|2020-09-30 ···················· ···················· ···················· ···················· ···················· ····················
Q 6806 hCoV-19/Norway/4673/2020|EPI_ISL_635066|2020-10-22 ···················· ···················· ···················· ···················· ···················· ····················
Q 6807 hCoV-19/Norway/4682/2020|EPI_ISL_635067|2020-10-22 ···················· ···················· ···················· ···················· ···················· ····················
Q 6808 hCoV-19/Norway/4695/2020|EPI_ISL_635068|2020-10-14 ···················· ···················· ···················· ···················· ···T················ ····················
Q 6809 hCoV-19/Norway/4697/2020|EPI_ISL_635069|2020-10-15 ···················· ···················· ···················· ···················· ···················· ····················
Q 6810 hCoV-19/Norway/4698/2020|EPI_ISL_635070|2020-10-15 ···················· ···················· ···················· ···················· ···················· ····················
Q 6811 hCoV-19/Norway/4701/2020|EPI_ISL_635071|2020-10-12 ···················· ···················· ···················· ···················· ···················· ····················
Q 6812 hCoV-19/Norway/4702/2020|EPI_ISL_635072|2020-10-12 ···················· ···················· ···················· ···················· ···················· ····················
Q 6813 hCoV-19/Norway/4745/2020|EPI_ISL_635073|2020-10-20 ···················· ···················· ···················· ···················· ···················· ····················
Q 6814 hCoV-19/Norway/4814/2020|EPI_ISL_635074|2020-10-26 ···················· ·A·················· ···················· ···················· ···················· ··A·················
Q 6815 hCoV-19/Norway/4817/2020|EPI_ISL_635075|2020-10-26 ···················· ···················· ···················· ···················· ···················· ····················
Q 6816 hCoV-19/Norway/4818/2020|EPI_ISL_635076|2020-10-26 ···················· ···················· ···················· ···················· ···················· ····················
Q 6817 hCoV-19/Norway/4852/2020|EPI_ISL_635077|2020-10-27 ···················· ···················· ···················· ···················· ···················· ····················
Q 6818 hCoV-19/Norway/4853/2020|EPI_ISL_635078|2020-10-27 ···················· ···················· ···················· ···················· ···················· ····················
Q 6819 hCoV-19/Norway/4963/2020|EPI_ISL_635079|2020-10-26 ···················· ···················· ···················· ···················· ···················· ····················
Q 6820 hCoV-19/Norway/4965/2020|EPI_ISL_635081|2020-10-26 ···················· ···················· ···················· ···················· ···················· ····················
Q 6821 hCoV-19/Norway/4993/2020|EPI_ISL_635085|2020-10-22 ···················· ···················· ···················· ···················· ···················· ····················
Q 6822 hCoV-19/Norway/5057/2020|EPI_ISL_635086|2020-10-29 ···················· ·A·················· ···················· ···················· ···················· ··A·················
Q 6823 hCoV-19/Norway/5060/2020|EPI_ISL_635087|2020-10-29 ···················· ·A·················· ···················· ···················· ···················· ··A·················
Q 6824 hCoV-19/Norway/4641/2020|EPI_ISL_635095|2020-10-04 ···················· ···················· ···················· ···················· ···················· ····················
Q 6825 hCoV-19/Norway/4056/2020|EPI_ISL_635096|2020-10-02 ···················· ···················· ···················· ···················· ···················· ····················
Q 6826 hCoV-19/Norway/4050/2020|EPI_ISL_635097|2020-10-03 ···················· ···················· ···················· ···················· ···················· ····················
Q 6827 hCoV-19/Norway/4613/2020|EPI_ISL_635098|2020-09-30 ···················· ···················· ···················· ···················· ···················· ····················
Q 6828 hCoV-19/Norway/3618/2020|EPI_ISL_635099|2020-09-24 ···················· ···················· ···················· ···················· ···················· ····················
Q 6829 hCoV-19/Jordan/AM-ALSR-3936/2020|EPI_ISL_636300|20 ···················· ···················· ···················· ···················· ···················· ····················
Q 6830 hCoV-19/Jordan/AM-ALSR-3937/2020|EPI_ISL_636301|20 ···················· ···················· ···················· ···················· ···················· ····················
Q 6831 hCoV-19/Jordan/AM-ALSR-3938/2020|EPI_ISL_636302|20 ···················· ···················· ···················· ···················· ···················· ····················
Q 6832 hCoV-19/Jordan/AM-ALSR-3939/2020|EPI_ISL_636303|20 ···················· ···················· ···················· ···················· ···················· ····················
Q 6833 hCoV-19/Jordan/AM-ALSR-3940/2020|EPI_ISL_636304|20 ···················· ···················· ···················· ···················· ···················· ····················
Q 6834 hCoV-19/Jordan/AM-ALSR-3941/2020|EPI_ISL_636305|20 ···················· ···················· ···················· ···················· ···················· ····················
Q 6835 hCoV-19/Jordan/AM-ALSR-3942/2020|EPI_ISL_636306|20 ···················· ···················· ···················· ···················· ···················· ····················
Q 6836 hCoV-19/Jordan/AM-ALSR-3943/2020|EPI_ISL_636307|20 ···················· ···················· ···················· ···················· ···················· ····················
Q 6837 hCoV-19/Jordan/AM-ALSR-3944/2020|EPI_ISL_636308|20 ···················· ···················· ···················· ···················· ···················· ····················
Q 6838 hCoV-19/Jordan/AM-ALSR-3945/2020|EPI_ISL_636309|20 ···················· ···················· ···················· ···················· ···················· ····················
Q 6839 hCoV-19/Jordan/AM-ALSR-3946/2020|EPI_ISL_636310|20 ···················· ···················· ···················· ···················· ···················· ····················
Q 6840 hCoV-19/Jordan/AM-ALSR-3947/2020|EPI_ISL_636311|20 ···················· ···················· ···················· ···················· ···················· ····················
Q 6841 hCoV-19/Jordan/AM-ALSR-3948/2020|EPI_ISL_636312|20 ···················· ···················· ···················· ···················· ···················· ····················
Q 6842 hCoV-19/Jordan/IR-ALSR-3949/2020|EPI_ISL_636313|20 ···················· ···················· ···················· ···················· ···················· ····················
Q 6843 hCoV-19/Jordan/AM-ALSR-3950/2020|EPI_ISL_636314|20 ···················· ···················· ···················· ···················· ···················· ····················
Q 6844 hCoV-19/Jordan/AM-ALSR-3951/2020|EPI_ISL_636315|20 ···················· ···················· ···················· ···················· ···················· ····················
Q 6845 hCoV-19/Jordan/AM-ALSR-3952/2020|EPI_ISL_636316|20 ···················· ···················· ···················· ···················· ···················· ····················
Q 6846 hCoV-19/Jordan/AM-ALSR-3954/2020|EPI_ISL_636317|20 ···················· ···················· ···················· ···················· ···················· ····················
Q 6847 hCoV-19/Jordan/AM-ALSR-3955/2020|EPI_ISL_636318|20 ···················· ···················· ···················· ···················· ···················· ····················

| | | | | | | | |
|---|---|---|---|---|---|---|---|
| Q | 6848 hCoV-19/Jordan/AM-ALSR-3956/2020\|EPI_ISL_636319\|20 | ···················· | ···················· | ····················· | ···················· | ···················· | ···················· |
| Q | 6849 hCoV-19/Jordan/AM-ALSR-3957/2020\|EPI_ISL_636320\|20 | ···················· | ···················· | ····················· | ···················· | ···················· | ···················· |
| Q | 6850 hCoV-19/Jordan/AM-ALSR-3958/2020\|EPI_ISL_636321\|20 | ···················· | ···················· | ····················· | ···················· | ···················· | ···················· |
| Q | 6851 hCoV-19/Jordan/AM-ALSR-3959/2020\|EPI_ISL_636322\|20 | ···················· | ···················· | ····················· | ···················· | ···················· | ···················· |
| Q | 6852 hCoV-19/Jordan/AM-ALSR-3960/2020\|EPI_ISL_636323\|20 | ···················· | ···················· | ····················· | ···················· | ···················· | ···················· |
| Q | 6853 hCoV-19/Jordan/AM-ALSR-3961/2020\|EPI_ISL_636324\|20 | ···················· | ···················· | ····················· | ···················· | ···················· | ···················· |
| Q | 6854 hCoV-19/Jordan/AM-ALSR-3962/2020\|EPI_ISL_636325\|20 | ···················· | ···················· | ····················· | ···················· | ···················· | ···················· |
| Q | 6855 hCoV-19/Jordan/AM-ALSR-3963/2020\|EPI_ISL_636326\|20 | ···················· | ·A·················· | ····················· | ···················· | ···················· | ··A················· |
| Q | 6856 hCoV-19/Jordan/AM-ALSR-3964/2020\|EPI_ISL_636327\|20 | ···················· | ·A·················· | ····················· | ···················· | ···················· | ··A················· |
| Q | 6857 hCoV-19/Jordan/AM-ALSR-3965/2020\|EPI_ISL_636328\|20 | ···················· | ···················· | ····················· | ···················· | ···················· | ···················· |
| Q | 6858 hCoV-19/Jordan/AM-ALSR-3966/2020\|EPI_ISL_636329\|20 | ···················· | ···················· | ····················· | ···················· | ·····T·············· | ···················· |
| Q | 6859 hCoV-19/Jordan/AM-ALSR-3967/2020\|EPI_ISL_636330\|20 | ···················· | ···················· | ····················· | ···················· | ·····T·············· | ···················· |
| Q | 6860 hCoV-19/Jordan/AM-ALSR-3968/2020\|EPI_ISL_636331\|20 | ···················· | ···················· | ····················· | ···················· | ···················· | ···················· |
| Q | 6861 hCoV-19/Jordan/AM-ALSR-3969/2020\|EPI_ISL_636332\|20 | ···················· | ···················· | ····················· | ···················· | ···················· | ···················· |
| Q | 6862 hCoV-19/Jordan/AM-ALSR-3970/2020\|EPI_ISL_636333\|20 | ···················· | ···················· | ····················· | ···················· | ···················· | ···················· |
| Q | 6863 hCoV-19/Jordan/AM-ALSR-3971/2020\|EPI_ISL_636334\|20 | ···················· | ···················· | ····················· | ···················· | ···················· | ···················· |
| Q | 6864 hCoV-19/Jordan/AM-ALSR-3972/2020\|EPI_ISL_636335\|20 | ···················· | ···················· | ····················· | ···················· | ···················· | ···················· |
| Q | 6865 hCoV-19/Jordan/AM-ALSR-3973/2020\|EPI_ISL_636336\|20 | ···················· | ···················· | ····················· | ···················· | ···················· | ···················· |
| Q | 6866 hCoV-19/Jordan/AM-ALSR-3974/2020\|EPI_ISL_636337\|20 | ···················· | ·A·················· | ····················· | ···················· | ···················· | ··A················· |
| Q | 6867 hCoV-19/Jordan/AM-ALSR-3975/2020\|EPI_ISL_636338\|20 | ···················· | ·A·················· | ····················· | ···················· | ···················· | ··A················· |
| Q | 6868 hCoV-19/Jordan/AM-ALSR-3976/2020\|EPI_ISL_636339\|20 | ···················· | ···················· | ····················· | ···················· | ···················· | ···················· |
| Q | 6869 hCoV-19/Jordan/AM-ALSR-3977/2020\|EPI_ISL_636340\|20 | ···················· | ···················· | ····················· | ···················· | ···················· | ···················· |
| Q | 6870 hCoV-19/Jordan/AM-ALSR-3978/2020\|EPI_ISL_636341\|20 | ···················· | ···················· | ····················· | ···················· | ···················· | ···················· |
| Q | 6871 hCoV-19/Jordan/AM-ALSR-3979/2020\|EPI_ISL_636342\|20 | ···················· | ···················· | ····················· | ···················· | ···················· | ···················· |
| Q | 6872 hCoV-19/Jordan/AM-ALSR-3980/2020\|EPI_ISL_636343\|20 | ···················· | ·A·················· | ····················· | ···················· | ···················· | ··A················· |
| Q | 6873 hCoV-19/Jordan/AM-ALSR-3984/2020\|EPI_ISL_636347\|20 | ···················· | ···················· | ····················· | ···················· | ···················· | ···················· |
| Q | 6874 hCoV-19/Jordan/AM-ALSR-3985/2020\|EPI_ISL_636348\|20 | ···················· | ···················· | ····················· | ···················· | ···················· | ···················· |
| Q | 6875 hCoV-19/Jordan/AM-ALSR-3986/2020\|EPI_ISL_636349\|20 | ···················· | ···················· | ····················· | ···················· | ···················· | ···················· |
| Q | 6876 hCoV-19/Jordan/AM-ALSR-3997/2020\|EPI_ISL_636360\|20 | ···················· | ···················· | ····················· | ···················· | ···················· | ···················· |
| Q | 6877 hCoV-19/Jordan/AM-ALSR-3999/2020\|EPI_ISL_636362\|20 | ···················· | ···················· | ····················· | ···················· | ···················· | ···················· |
| Q | 6878 hCoV-19/Jordan/AM-ALSR-4000/2020\|EPI_ISL_636363\|20 | ···················· | ···················· | ····················· | ···················· | ···················· | ···················· |
| Q | 6879 hCoV-19/Jordan/AM-ALSR-4001/2020\|EPI_ISL_636364\|20 | ···················· | ···················· | ····················· | ···················· | ···················· | ···················· |
| Q | 6880 hCoV-19/Jordan/AM-ALSR-4002/2020\|EPI_ISL_636365\|20 | ···················· | ···················· | ····················· | ···················· | ···················· | ···················· |
| Q | 6881 hCoV-19/Jordan/AM-ALSR-4003/2020\|EPI_ISL_636366\|20 | ···················· | ···················· | ····················· | ···················· | ···················· | ···················· |
| Q | 6882 hCoV-19/Jordan/AM-ALSR-4004/2020\|EPI_ISL_636367\|20 | ···················· | ···················· | ····················· | ···················· | ···················· | ···················· |
| Q | 6883 hCoV-19/Jordan/AM-ALSR-4005/2020\|EPI_ISL_636368\|20 | ···················· | ···················· | ····················· | ···················· | ···················· | ···················· |
| Q | 6884 hCoV-19/Jordan/AM-ALSR-4006/2020\|EPI_ISL_636369\|20 | ···················· | ···················· | ····················· | ···················· | ···················· | ···················· |
| Q | 6885 hCoV-19/Jordan/AM-ALSR-4007/2020\|EPI_ISL_636370\|20 | ···················· | ·A·················· | ····················· | ···················· | ···················· | ··A················· |
| Q | 6886 hCoV-19/Jordan/AM-ALSR-4008/2020\|EPI_ISL_636371\|20 | ···················· | ···················· | ····················· | ···················· | ···················· | ···················· |
| Q | 6887 hCoV-19/Jordan/AM-ALSR-4009/2020\|EPI_ISL_636372\|20 | ···················· | ···················· | ····················· | ···················· | ···················· | ···················· |
| Q | 6888 hCoV-19/Jordan/AM-ALSR-4010/2020\|EPI_ISL_636373\|20 | ···················· | ···················· | ····················· | ···················· | ···················· | ···················· |
| Q | 6889 hCoV-19/Jordan/AM-ALSR-4011/2020\|EPI_ISL_636374\|20 | ···················· | ···················· | ····················· | ···················· | ···················· | ···················· |
| Q | 6890 hCoV-19/Jordan/AM-ALSR-4012/2020\|EPI_ISL_636375\|20 | ···················· | ···················· | ····················· | ···················· | ···················· | ···················· |
| Q | 6891 hCoV-19/Jordan/AM-ALSR-4013/2020\|EPI_ISL_636376\|20 | ···················· | ···················· | ····················· | ···················· | ···················· | ···················· |
| Q | 6892 hCoV-19/Jordan/AM-ALSR-4014/2020\|EPI_ISL_636377\|20 | ···················· | ···················· | ····················· | ···················· | ···················· | ···················· |
| Q | 6893 hCoV-19/Jordan/AM-ALSR-4015/2020\|EPI_ISL_636378\|20 | ···················· | ···················· | ····················· | ···················· | ···················· | ···················· |
| Q | 6894 hCoV-19/Jordan/AM-ALSR-4016/2020\|EPI_ISL_636379\|20 | ···················· | ···················· | ····················· | ···················· | ···················· | ···················· |
| Q | 6895 hCoV-19/Jordan/AM-ALSR-4017/2020\|EPI_ISL_636380\|20 | ···················· | ···················· | ····················· | ···················· | ···················· | ···················· |
| Q | 6896 hCoV-19/Jordan/AM-ALSR-4020/2020\|EPI_ISL_636381\|20 | ···················· | ···················· | ····················· | ···················· | ···················· | ···················· |
| Q | 6897 hCoV-19/Jordan/AM-ALSR-4021/2020\|EPI_ISL_636382\|20 | ···················· | ···················· | ····················· | ···················· | ···················· | ···················· |
| Q | 6898 hCoV-19/Jordan/AM-ALSR-4022/2020\|EPI_ISL_636383\|20 | ···················· | ···················· | ····················· | ···················· | ···················· | ···················· |
| Q | 6899 hCoV-19/Jordan/AM-ALSR-4023/2020\|EPI_ISL_636384\|20 | ···················· | ···················· | ····················· | ···················· | ···················· | ···················· |
| Q | 6900 hCoV-19/Jordan/AM-ALSR-4024/2020\|EPI_ISL_636385\|20 | ···················· | ···················· | ····················· | ···················· | ···················· | ···················· |
| Q | 6901 hCoV-19/Jordan/AM-ALSR-4025/2020\|EPI_ISL_636386\|20 | ···················· | ···················· | ····················· | ···················· | ···················· | ···················· |
| Q | 6902 hCoV-19/Jordan/AM-ALSR-4027/2020\|EPI_ISL_636388\|20 | ···················· | ···················· | ····················· | ···················· | ···················· | ···················· |
| Q | 6903 hCoV-19/Jordan/AM-ALSR-4028/2020\|EPI_ISL_636389\|20 | ···················· | ···················· | ····················· | ···················· | ···················· | ···················· |
| Q | 6904 hCoV-19/Jordan/AM-ALSR-4031/2020\|EPI_ISL_636391\|20 | ···················· | ···················· | ····················· | ···················· | ···················· | ···················· |
| Q | 6905 hCoV-19/Jordan/AM-ALSR-4032/2020\|EPI_ISL_636392\|20 | ···················· | ···················· | ····················· | ···················· | ···················· | ···················· |
| Q | 6906 hCoV-19/Jordan/AM-ALSR-4033/2020\|EPI_ISL_636393\|20 | ···················· | ···················· | ····················· | ···················· | ···················· | ···················· |
| Q | 6907 hCoV-19/Jordan/AM-ALSR-4034/2020\|EPI_ISL_636394\|20 | ···················· | ···················· | ····················· | ···················· | ···················· | ···················· |
| Q | 6908 hCoV-19/Jordan/AM-ALSR-4035/2020\|EPI_ISL_636395\|20 | ···················· | ···················· | ····················· | ···················· | ···················· | ···················· |
| Q | 6909 hCoV-19/Jordan/AM-ALSR-4036/2020\|EPI_ISL_636396\|20 | ···················· | ···················· | ····················· | ···················· | ···················· | ···················· |
| Q | 6910 hCoV-19/Jordan/AM-ALSR-4037/2020\|EPI_ISL_636397\|20 | ···················· | ···················· | ····················· | ···················· | ···················· | ···················· |
| Q | 6911 hCoV-19/Jordan/AM-ALSR-4038/2020\|EPI_ISL_636398\|20 | ···················· | ···················· | ····················· | ···················· | ···················· | ···················· |
| Q | 6912 hCoV-19/Jordan/AM-ALSR-4039/2020\|EPI_ISL_636399\|20 | ···················· | ·A·················· | ····················· | ···················· | ···················· | ··A················· |
| Q | 6913 hCoV-19/France/ARA-SC060/2020\|EPI_ISL_629101\|2020- | ···················· | ···················· | ····················· | ···················· | ···················· | ···················· |
| Q | 6914 hCoV-19/France/ARA-SC064/2020\|EPI_ISL_629102\|2020- | ···················· | ···················· | ····················· | ···················· | ···················· | ···················· |
| Q | 6915 hCoV-19/France/ARA-SC066/2020\|EPI_ISL_629103\|2020- | ···················· | ···················· | ····················· | ···················· | ···················· | ···················· |
| Q | 6917 hCoV-19/France/ARA-SC070/2020\|EPI_ISL_629105\|2020- | ···················· | ···················· | ····················· | ···················· | ···················· | ···················· |
| Q | 6918 hCoV-19/France/ARA-SC073/2020\|EPI_ISL_629106\|2020- | ···················· | ···················· | ····················· | ···················· | ···················· | ···················· |
| Q | 6919 hCoV-19/France/ARA-SC075/2020\|EPI_ISL_629107\|2020- | ···················· | ···················· | ····················· | ···················· | ···················· | ···················· |
| Q | 6920 hCoV-19/France/ARA-SC078/2020\|EPI_ISL_629108\|2020- | ···················· | ···················· | ····················· | ···················· | ···················· | ···················· |
| Q | 6921 hCoV-19/England/MILK-ACC394/2020\|EPI_ISL_629109\|20 | ···················· | ···················· | ····················· | ···················· | ···················· | ···················· |
| Q | 6922 hCoV-19/England/MILK-ACC622/2020\|EPI_ISL_629110\|20 | ···················· | ···················· | ····················· | ···················· | ···················· | ···················· |
| Q | 6923 hCoV-19/England/MILK-AC94D0/2020\|EPI_ISL_629111\|20 | ···················· | ·A·················· | ····················· | ···················· | ···················· | ··A················· |
| Q | 6924 hCoV-19/England/MILK-ACC19A/2020\|EPI_ISL_629112\|20 | ···················· | ···················· | ····················· | ···················· | ···················· | ···················· |
| Q | 6925 hCoV-19/England/MILK-ACE365/2020\|EPI_ISL_629113\|20 | ···················· | ···················· | ····················· | ···················· | ···················· | ···················· |
| Q | 6926 hCoV-19/England/MILK-ACBC9A/2020\|EPI_ISL_629114\|20 | ···················· | ···················· | ····················· | ···················· | ···················· | ···················· |
| Q | 6927 hCoV-19/England/MILK-AC85DE/2020\|EPI_ISL_629115\|20 | ···················· | ·A·················· | ····················· | ···················· | ···················· | ···················· |
| Q | 6928 hCoV-19/England/MILK-AC85FC/2020\|EPI_ISL_629116\|20 | ···················· | ···················· | ····················· | ···················· | ···················· | ···················· |

Q 6929 hCoV-19/England/MILK-ACBD00/2020|EPI_ISL_629117|20 ···················· ···················· ····················· ···················· ······T············· ····················

Q 6930 hCoV-19/England/MILK-ACBEFE/2020|EPI_ISL_629118|20 ···················· ···················· ····················· ···················· ···················· ····················

Q 6931 hCoV-19/England/MILK-ACC09D/2020|EPI_ISL_629119|20 ···················· ···················· ···········Y········· ···················· ···················· ····················

Q 6932 hCoV-19/England/MILK-ACC8E0/2020|EPI_ISL_629121|20 ···················· ···················· ····················· ···················· ···················· ····················

Q 6933 hCoV-19/England/MILK-ACC8D1/2020|EPI_ISL_629122|20 ···················· ···················· ····················· ···················· ···················· ····················

Q 6934 hCoV-19/England/MILK-ACE893/2020|EPI_ISL_629123|20 ···················· ···················· ····················· ···················· ···················· ····················

Q 6935 hCoV-19/England/MILK-ACCA71/2020|EPI_ISL_629124|20 ···················· ···················· ····················· ···················· ···················· ····················

Q 6936 hCoV-19/England/MILK-ACE4CC/2020|EPI_ISL_629125|20 ···················· ···················· ····················· ···················· ···················· ····················

Q 6937 hCoV-19/England/MILK-AC9537/2020|EPI_ISL_629126|20 ···················· ···················· ····················· ···················· ···················· ····················

Q 6938 hCoV-19/England/MILK-ACC25B/2020|EPI_ISL_629127|20 ···················· ···················· ····················· ···················· ···················· ····················

Q 6939 hCoV-19/England/MILK-ACE505/2020|EPI_ISL_629128|20 ···················· ···················· ····················· ···················· ···················· ····················

Q 6940 hCoV-19/England/MILK-ACE426/2020|EPI_ISL_629129|20 ···················· ···················· ····················· ···················· ···················· ····················

Q 6941 hCoV-19/England/MILK-ACBCA9/2020|EPI_ISL_629130|20 ···················· ·A·······A·········· ····················· ···················· ···················· ····················

Q 6942 hCoV-19/England/MILK-AC837A/2020|EPI_ISL_629131|20 ···················· ···················· ····················· ···················· ····T··············· ····················

Q 6943 hCoV-19/England/MILK-ACBCC7/2020|EPI_ISL_629132|20 ···················· ···················· ····················· ···················· ···················· ····················

Q 6944 hCoV-19/England/MILK-ACBD79/2020|EPI_ISL_629133|20 ···················· ···················· ····················· ···················· ···················· ····················

Q 6945 hCoV-19/England/MILK-AC82E6/2020|EPI_ISL_629134|20 ···················· ···················· ····················· ···················· ···················· ····················

Q 6946 hCoV-19/England/MILK-ACE909/2020|EPI_ISL_629135|20 ···················· ···················· ····················· ···················· ···················· ····················

Q 6947 hCoV-19/England/MILK-ACE49F/2020|EPI_ISL_629136|20 ···················· ···················· ····················· ···················· ···················· ····················

Q 6948 hCoV-19/England/MILK-ACBD88/2020|EPI_ISL_629139|20 ···················· ···················· ····················· ···················· ···················· ····················

Q 6949 hCoV-19/England/MILK-ACC22E/2020|EPI_ISL_629142|20 ···················· ···················· ····················· ···················· ···················· ····················

Q 6950 hCoV-19/England/MILK-ACE972/2020|EPI_ISL_629143|20 ···················· ···················· ····················· ···················· ···················· ····················

Q 6951 hCoV-19/England/MILK-ACED49/2020|EPI_ISL_629144|20 ···················· ·A·················· ····················· ···················· ···················· ···A················

Q 6952 hCoV-19/England/MILK-ACE3FC/2020|EPI_ISL_629145|20 ···················· ···················· ····················· ···················· ···················· ····················

Q 6953 hCoV-19/England/MILK-ACBFA0/2020|EPI_ISL_629146|20 ···················· ···················· ····················· ···················· ······T············· ····················

Q 6954 hCoV-19/England/MILK-ACE5F6/2020|EPI_ISL_629147|20 ···················· ···················· ····················· ···················· ···················· ····················

Q 6955 hCoV-19/England/MILK-ACBD1F/2020|EPI_ISL_629148|20 ···················· ···················· ····················· ···················· ···················· ····················

Q 6956 hCoV-19/England/MILK-AC9519/2020|EPI_ISL_629149|20 ···················· ···················· ····················· ···················· ···················· ····················

Q 6957 hCoV-19/England/MILK-ACBE0D/2020|EPI_ISL_629150|20 ···················· ···················· ····················· ···················· ···················· ····················

Q 6958 hCoV-19/England/MILK-ACE4BD/2020|EPI_ISL_629151|20 ···················· ···················· ····················· ···················· ···················· ····················

Q 6959 hCoV-19/England/MILK-AC83B6/2020|EPI_ISL_629152|20 ···················· ···················· ····················· ···················· ···················· ····················

Q 6960 hCoV-19/England/MILK-ACE3ED/2020|EPI_ISL_629153|20 ···················· ···················· ····················· ···················· ···················· ····················

Q 6961 hCoV-19/England/MILK-AC7D8C/2020|EPI_ISL_629154|20 ···················· ···················· ····················· ···················· ···················· ····················

Q 6962 hCoV-19/England/MILK-AC928B/2020|EPI_ISL_629155|20 ···················· ···················· ····················· ···················· ···················· ····················

Q 6963 hCoV-19/England/MILK-ACC76B/2020|EPI_ISL_629156|20 ···················· ···················· ····················· ···················· ···················· ····················

Q 6964 hCoV-19/England/MILK-AC9616/2020|EPI_ISL_629157|20 ···················· ···················· ····················· ···················· ···················· ····················

Q 6965 hCoV-19/England/MILK-ACC8FF/2020|EPI_ISL_629158|20 ···················· ·A·················· ····················· ···················· ···················· ···A················

Q 6966 hCoV-19/England/MILK-AC9652/2020|EPI_ISL_629159|20 ···················· ···················· ····················· ···················· ···················· ····················

Q 6967 hCoV-19/England/MILK-ACE374/2020|EPI_ISL_629160|20 ···················· ···················· ····················· ···················· ···················· ····················

Q 6968 hCoV-19/England/MILK-ACCA17/2020|EPI_ISL_629161|20 ···················· ·A·················· ····················· ···················· ···················· ···A················

Q 6969 hCoV-19/England/MILK-ACE329/2020|EPI_ISL_629162|20 ···················· ···················· ····················· ···················· ···················· ····················

Q 6970 hCoV-19/England/MILK-AC869F/2020|EPI_ISL_629163|20 ···················· ···················· ····················· ···················· ···················· ····················

Q 6971 hCoV-19/England/MILK-AC9607/2020|EPI_ISL_629164|20 ···················· ···················· ····················· ···················· ···················· ····················

Q 6972 hCoV-19/England/MILK-ACECA6/2020|EPI_ISL_629165|20 ···················· ···················· ····················· ···················· ···················· ····················

Q 6973 hCoV-19/England/MILK-ACE356/2020|EPI_ISL_629166|20 ···················· ···················· ····················· ···················· ···················· ····················

Q 6974 hCoV-19/England/MILK-ACC1B8/2020|EPI_ISL_629167|20 ···················· ···················· ····················· ···················· ······T············· ···············A····

Q 6975 hCoV-19/England/MILK-ACE884/2020|EPI_ISL_629168|20 ···················· ···················· ····················· ···················· ···················· ····················

Q 6976 hCoV-19/England/MILK-AC8671/2020|EPI_ISL_629169|20 ···················· ···················· ····················· ···················· ···················· ····················

Q 6977 hCoV-19/England/MILK-ACE286/2020|EPI_ISL_629180|20 ···················· ···················· ····················· ···················· ···················· ····················

Q 6978 hCoV-19/England/MILK-ACBF19/2020|EPI_ISL_629181|20 ···················· ···················· ····················· ···················· ······T············· ····················

Q 6979 hCoV-19/England/MILK-ACBCE5/2020|EPI_ISL_629182|20 ···················· ···················· ····················· ···················· ···················· ····················

Q 6980 hCoV-19/England/MILK-ACBD3D/2020|EPI_ISL_629183|20 ···················· ···················· ····················· ···················· ···················· ····················

Q 6981 hCoV-19/England/MILK-ACED2B/2020|EPI_ISL_629184|20 ···················· ···················· ····················· ···················· ···················· ····················

Q 6982 hCoV-19/England/MILK-AC92B8/2020|EPI_ISL_629185|20 ···················· ···················· ····················· ···················· ···················· ····················

Q 6983 hCoV-19/England/MILK-AC72BA/2020|EPI_ISL_630100|20 ···················· ···················· ····················· ···················· ···················· ····················

Q 6984 hCoV-19/England/MILK-AC93D3/2020|EPI_ISL_630101|20 ···················· ···················· ····················· ···················· ···················· ····················

Q 6985 hCoV-19/England/MILK-AC9397/2020|EPI_ISL_630102|20 ···················· ···················· ····················· ···················· ···················· ····················

Q 6986 hCoV-19/England/MILK-ACE1B6/2020|EPI_ISL_630103|20 ···················· ···················· ····················· ···················· ······T············· ····················

Q 6987 hCoV-19/England/MILK-AC76DC/2020|EPI_ISL_630104|20 ···················· ···················· ····················· ···················· ···················· ····················

Q 6988 hCoV-19/England/MILK-ACDEB0/2020|EPI_ISL_630105|20 ···················· ···················· ····················· ···················· ···················· ····················

Q 6989 hCoV-19/England/MILK-AC9A56/2020|EPI_ISL_630106|20 ···················· ···················· ····················· ···················· ···················· ····················

Q 6990 hCoV-19/England/MILK-ACCC99/2020|EPI_ISL_630107|20 ···················· ···················· ····················· ···················· ······T············· ····················

Q 6991 hCoV-19/England/MILK-ACDBAA/2020|EPI_ISL_630108|20 ···················· ···················· ····················· ···················· ···················· ····················

Q 6992 hCoV-19/England/MILK-ACDDC2/2020|EPI_ISL_630111|20 ···················· ·A·················· ····················· ···················· ···················· ···A················

Q 6993 hCoV-19/England/MILK-ACCCF3/2020|EPI_ISL_630112|20 ···················· ···················· ····················· ···················· ···················· ····················

Q 6994 hCoV-19/England/MILK-AC9106/2020|EPI_ISL_630113|20 ···················· ···················· ····················· ···················· ······T············· ····················

Q 6995 hCoV-19/England/MILK-ACDF71/2020|EPI_ISL_630115|20 ···················· ···················· ····················· ···················· ···················· ····················

Q 6996 hCoV-19/England/MILK-ACCC6C/2020|EPI_ISL_630116|20 ···················· ···················· ····················· ···················· ···················· ····················

Q 6997 hCoV-19/England/MILK-AC941C/2020|EPI_ISL_630117|20 ···················· ···················· ····················· ···················· ···················· ····················

Q 6998 hCoV-19/England/MILK-ACD876/2020|EPI_ISL_630118|20 ···················· ···················· ····················· ···················· ···················· ····················

Q 6999 hCoV-19/England/MILK-ACD9A0/2020|EPI_ISL_630119|20 ···················· ···················· ····················· ···················· ···················· ····················

Q 7000 hCoV-19/England/MILK-AC76EB/2020|EPI_ISL_630120|20 ···················· ···················· ····················· ···················· ···················· ····················

Q 7001 hCoV-19/England/MILK-ACD2C3/2020|EPI_ISL_630121|20 ···················· ···················· ····················· ···················· ···················· ····················

Q 7002 hCoV-19/England/MILK-ACCE39/2020|EPI_ISL_630122|20 ···················· ···················· ····················· ···················· ···················· ····················

Q 7003 hCoV-19/England/MILK-AC750C/2020|EPI_ISL_630123|20 ···················· ···················· ····················· ···················· ···················· ····················

Q 7004 hCoV-19/England/MILK-ACCB14/2020|EPI_ISL_630124|20 ···················· ···················· ····················· ···················· ···················· ····················

Q 7005 hCoV-19/England/MILK-AC779D/2020|EPI_ISL_630125|20 ···················· ···················· ····················· ···················· ···················· ····················

Q 7006 hCoV-19/England/MILK-ACD8FE/2020|EPI_ISL_630126|20 ···················· ···················· ····················· ···················· ···················· ····················

Q 7007 hCoV-19/England/MILK-ACE031/2020|EPI_ISL_630128|20 ···················· ···················· ····················· ···················· ···················· ····················

Q 7008 hCoV-19/England/MILK-ACE15C/2020|EPI_ISL_630137|20 ···················· ···················· ····················· ···················· ······T············· ····················

| | | | | | | | |
|---|---|---|---|---|---|---|---|
| Q | 7009 hCoV-19/England/MILK-AC72AB/2020\|EPI_ISL_630138\|20 | ···················· | ···················· | ····················· | ···················· | ···················· | ···················· |
| Q | 7010 hCoV-19/England/MILK-AC7715/2020\|EPI_ISL_630139\|20 | ···················· | ···················· | ····················· | ···················· | ···················· | ···················· |
| Q | 7011 hCoV-19/England/MILK-ACE09B/2020\|EPI_ISL_630140\|20 | ···················· | ···················· | ····················· | ···················· | ···················· | ···················· |
| Q | 7012 hCoV-19/England/MILK-AC7681/2020\|EPI_ISL_630141\|20 | ···················· | ···················· | ····················· | ···················· | ···················· | ···················· |
| Q | 7013 hCoV-19/England/MILK-ACDEFC/2020\|EPI_ISL_630142\|20 | ···················· | ···················· | ····················· | ···················· | ···················· | ···················· |
| Q | 7014 hCoV-19/England/MILK-ACE0C8/2020\|EPI_ISL_630143\|20 | ···················· | ···················· | ····················· | ···················· | ····T··············· | ···················· |
| Q | 7015 hCoV-19/England/MILK-ACD973/2020\|EPI_ISL_630144\|20 | ···················· | ·A·······A·········· | ····················· | ···················· | ···················· | ···················· |
| Q | 7016 hCoV-19/England/MILK-AC7584/2020\|EPI_ISL_630145\|20 | ···················· | ···················· | ····················· | ···················· | ···················· | ···················· |
| Q | 7017 hCoV-19/England/MILK-ACD2A5/2020\|EPI_ISL_630146\|20 | ···················· | ···················· | ····················· | ···················· | ···················· | ···················· |
| Q | 7018 hCoV-19/England/MILK-AC9A65/2020\|EPI_ISL_630148\|20 | ···················· | ···················· | ····················· | ···················· | ···················· | ···················· |
| Q | 7019 hCoV-19/England/MILK-AC74D2/2020\|EPI_ISL_630149\|20 | ···················· | ···················· | ····················· | ···················· | ···················· | ···················· |
| Q | 7020 hCoV-19/England/MILK-ACD023/2020\|EPI_ISL_630150\|20 | ···················· | ···················· | ····················· | ···················· | ···················· | ···················· |
| Q | 7021 hCoV-19/England/MILK-ACCF54/2020\|EPI_ISL_630152\|20 | ···················· | ···················· | ····················· | ···················· | ···················· | ···················· |
| Q | 7022 hCoV-19/England/MILK-AC735D/2020\|EPI_ISL_630153\|20 | ···················· | ···················· | ····················· | ···················· | ···················· | ···················· |
| Q | 7023 hCoV-19/England/MILK-ACCCB7/2020\|EPI_ISL_630154\|20 | ···················· | ···················· | ····················· | ···················· | ···················· | ···················· |
| Q | 7024 hCoV-19/England/MILK-ACE1D4/2020\|EPI_ISL_630155\|20 | ···················· | ···················· | ····················· | ···················· | ···················· | ···················· |
| Q | 7025 hCoV-19/England/MILK-ACCF90/2020\|EPI_ISL_630156\|20 | ···················· | ···················· | ····················· | ···················· | ···················· | ···················· |
| Q | 7026 hCoV-19/England/MILK-ACDD4A/2020\|EPI_ISL_630157\|20 | ···················· | ···················· | ····················· | ···················· | ···················· | ···················· |
| Q | 7027 hCoV-19/England/MILK-AC777F/2020\|EPI_ISL_630158\|20 | ···················· | ···················· | ····················· | ···················· | ···················· | ···················· |
| Q | 7028 hCoV-19/England/MILK-ACE05F/2020\|EPI_ISL_630171\|20 | ···················· | ···················· | ····················· | ···················· | ···················· | ···················· |
| Q | 7029 hCoV-19/England/MILK-ACD0BA/2020\|EPI_ISL_630172\|20 | ···················· | ···················· | ····················· | ···················· | ······T············· | ···················· |
| Q | 7030 hCoV-19/England/MILK-AC7539/2020\|EPI_ISL_630173\|20 | ···················· | ···················· | ····················· | ···················· | ···················· | ···················· |
| Q | 7031 hCoV-19/England/MILK-AC7706/2020\|EPI_ISL_630174\|20 | ···················· | ···················· | ····················· | ···················· | ···················· | ···················· |
| Q | 7032 hCoV-19/England/MILK-ACD2B4/2020\|EPI_ISL_630176\|20 | ···················· | ···················· | ····················· | ···················· | ···················· | ···················· |
| Q | 7033 hCoV-19/England/MILK-ACDFAE/2020\|EPI_ISL_630177\|20 | ···················· | ···················· | ····················· | ···················· | ···················· | ···················· |
| Q | 7034 hCoV-19/England/MILK-ACD0E7/2020\|EPI_ISL_630178\|20 | ···················· | ···················· | ····················· | ···················· | ···················· | ···················· |
| Q | 7055 hCoV-19/USA/WI-WSLH-200659/2020\|EPI_ISL_631425\|202 | ···················· | ···················· | ····················· | ···················· | ···················· | ···················· |
| Q | 7056 hCoV-19/USA/WI-WSLH-200660/2020\|EPI_ISL_631426\|202 | ···················· | ···················· | ····················· | ···················· | ···················· | ···················· |
| Q | 7063 hCoV-19/USA/WI-WSLH-200675/2020\|EPI_ISL_631433\|202 | ···················· | ···················· | ····················· | ···················· | ···················· | ···················· |
| Q | 7078 hCoV-19/USA/WI-WSLH-200691/2020\|EPI_ISL_631448\|202 | ···················· | ···················· | ····················· | ···················· | ···················· | ···················· |
| Q | 7084 hCoV-19/USA/WI-WSLH-200697/2020\|EPI_ISL_631454\|202 | ···················· | ···················· | ····················· | ···················· | ···················· | ···················· |
| Q | 7099 hCoV-19/USA/WI-WSLH-200722/2020\|EPI_ISL_631478\|202 | ···················· | ·A·················· | ····················· | ···················· | ···················· | ···················· |
| Q | 7100 hCoV-19/USA/WI-WSLH-200723/2020\|EPI_ISL_631479\|202 | ···················· | ·A·················· | ····················· | ···················· | ···················· | ···················· |
| Q | 7101 hCoV-19/USA/WI-WSLH-200724/2020\|EPI_ISL_631480\|202 | ···················· | ···················· | ····················· | ···················· | ···················· | ···················· |
| Q | 7102 hCoV-19/USA/WI-WSLH-200725/2020\|EPI_ISL_631481\|202 | ···················· | ···················· | ····················· | ···················· | ···················· | ···················· |
| Q | 7104 hCoV-19/USA/WI-WSLH-200727/2020\|EPI_ISL_631483\|202 | ···················· | ···················· | ····················· | ···················· | ···················· | ···················· |
| Q | 7105 hCoV-19/USA/WI-WSLH-200728/2020\|EPI_ISL_631484\|202 | ···················· | ···················· | ····················· | ···················· | ···················· | ···················· |
| Q | 7106 hCoV-19/USA/WI-WSLH-200729/2020\|EPI_ISL_631485\|202 | ···················· | ···················· | ····················· | ···················· | ···················· | ···················· |
| Q | 7107 hCoV-19/USA/WI-WSLH-200730/2020\|EPI_ISL_631486\|202 | ···················· | ···················· | ····················· | ···················· | ···················· | ···················· |
| Q | 7111 hCoV-19/Netherlands/UT-EMC-140/2020\|EPI_ISL_632733 | ···················· | ···················· | ····················· | ···················· | ······T············· | ···················· |
| Q | 7112 hCoV-19/Netherlands/ZH-EMC-852/2020\|EPI_ISL_632734 | ···················· | ···················· | ····················· | ···················· | ···················· | ···················· |
| Q | 7113 hCoV-19/Netherlands/ZH-EMC-853/2020\|EPI_ISL_632735 | ···················· | ···················· | ····················· | ···················· | ···················· | ···················· |
| Q | 7114 hCoV-19/Netherlands/ZH-EMC-854/2020\|EPI_ISL_632736 | ···················· | ·A·················· | ····················· | ···················· | ···················· | ···A················ |
| Q | 7115 hCoV-19/Netherlands/ZH-EMC-855/2020\|EPI_ISL_632737 | ···················· | ···················· | ····················· | ···················· | ···················· | ···················· |
| Q | 7116 hCoV-19/Netherlands/ZH-EMC-857/2020\|EPI_ISL_632738 | ···················· | ···················· | ····················· | ···················· | ······T············· | ···················· |
| Q | 7117 hCoV-19/Netherlands/GE-EMC-340/2020\|EPI_ISL_632739 | ···················· | ···················· | ····················· | ···················· | ···················· | ···················· |
| Q | 7118 hCoV-19/Netherlands/FL-EMC-36/2020\|EPI_ISL_632740\| | ···················· | ···················· | ····················· | ···················· | ···················· | ···················· |
| Q | 7119 hCoV-19/Netherlands/OV-EMC-83/2020\|EPI_ISL_632741\| | ···················· | ···················· | ····················· | ···················· | ···················· | ···················· |
| Q | 7120 hCoV-19/Netherlands/GE-EMC-342/2020\|EPI_ISL_632742 | ···················· | ···················· | ····················· | ···················· | ···················· | ···················· |
| Q | 7121 hCoV-19/Netherlands/GE-EMC-344/2020\|EPI_ISL_632743 | ···················· | ···················· | ····················· | ···················· | ···················· | ···················· |
| Q | 7122 hCoV-19/Netherlands/FL-EMC-37/2020\|EPI_ISL_632744\| | ···················· | ···················· | ····················· | ···················· | ···················· | ···················· |
| Q | 7123 hCoV-19/Netherlands/GE-EMC-346/2020\|EPI_ISL_632745 | ···················· | ···················· | ····················· | ···················· | ···················· | ···················· |
| Q | 7124 hCoV-19/Netherlands/ZH-EMC-860/2020\|EPI_ISL_632746 | ···················· | ···················· | ····················· | ···················· | ···················· | ···················· |
| Q | 7125 hCoV-19/Netherlands/ZH-EMC-862/2020\|EPI_ISL_632747 | ···················· | ···················· | ····················· | ···················· | ···················· | ···················· |
| Q | 7126 hCoV-19/Netherlands/ZH-EMC-863/2020\|EPI_ISL_632748 | ···················· | ···················· | ····················· | ···················· | ···················· | ···················· |
| Q | 7127 hCoV-19/Netherlands/ZH-EMC-864/2020\|EPI_ISL_632749 | ···················· | ···················· | ····················· | ···················· | ···················· | ···················· |
| Q | 7128 hCoV-19/Netherlands/ZH-EMC-865/2020\|EPI_ISL_632750 | ···················· | ···················· | ····················· | ···················· | ······T············· | ···················· |
| Q | 7129 hCoV-19/Netherlands/ZH-EMC-867/2020\|EPI_ISL_632751 | ···················· | ···················· | ····················· | ···················· | ···················· | ···················· |
| Q | 7130 hCoV-19/Netherlands/ZH-EMC-869/2020\|EPI_ISL_632752 | ···················· | ···················· | ····················· | ···················· | ······T············· | ···················· |
| Q | 7131 hCoV-19/Netherlands/ZH-EMC-870/2020\|EPI_ISL_632753 | ···················· | ···················· | ····················· | ···················· | ···················· | ···················· |
| Q | 7132 hCoV-19/Netherlands/ZH-EMC-872/2020\|EPI_ISL_632754 | ···················· | ···················· | ····················· | ···················· | ···················· | ···················· |
| Q | 7133 hCoV-19/Netherlands/ZH-EMC-874/2020\|EPI_ISL_632755 | ···················· | ···················· | ····················· | ···················· | ···················· | ···················· |
| Q | 7134 hCoV-19/Netherlands/ZH-EMC-876/2020\|EPI_ISL_632756 | ···················· | ···················· | ····················· | ···················· | ···················· | ···················· |
| Q | 7135 hCoV-19/Netherlands/ZH-EMC-877/2020\|EPI_ISL_632757 | ···················· | ···················· | ····················· | ···················· | ······T············· | ···················· |
| Q | 7136 hCoV-19/Netherlands/ZH-EMC-878/2020\|EPI_ISL_632758 | ···················· | ···················· | ····················· | ···················· | ···················· | ···················· |
| Q | 7137 hCoV-19/Netherlands/ZH-EMC-879/2020\|EPI_ISL_632759 | ···················· | ···················· | ····················· | ···················· | ···················· | ···················· |
| Q | 7138 hCoV-19/Netherlands/ZH-EMC-880/2020\|EPI_ISL_632760 | ···················· | ···················· | ····················· | ···················· | ···················· | ···················· |
| Q | 7139 hCoV-19/Netherlands/LI-EMC-100/2020\|EPI_ISL_632761 | ···················· | ···················· | ····················· | ···················· | ···················· | ···················· |
| Q | 7140 hCoV-19/Netherlands/ZH-EMC-883/2020\|EPI_ISL_632762 | ···················· | ···················· | ····················· | ···················· | ···················· | ···················· |
| Q | 7141 hCoV-19/Netherlands/ZH-EMC-884/2020\|EPI_ISL_632763 | ···················· | ···················· | ····················· | ···················· | ···················· | ···················· |
| Q | 7142 hCoV-19/Netherlands/GE-EMC-348/2020\|EPI_ISL_632764 | ···················· | ···················· | ····················· | ···················· | ···················· | ···················· |
| Q | 7143 hCoV-19/Netherlands/ZH-EMC-885/2020\|EPI_ISL_632765 | ···················· | ·A·················· | ····················· | ···················· | ···················· | ···A················ |
| Q | 7144 hCoV-19/Netherlands/ZH-EMC-886/2020\|EPI_ISL_632766 | ···················· | ···················· | ····················· | ···················· | ······T············· | ···················· |
| Q | 7145 hCoV-19/Netherlands/ZH-EMC-888/2020\|EPI_ISL_632767 | ···················· | ···················· | ···T················· | ···················· | ···················· | ···················· |
| Q | 7146 hCoV-19/Netherlands/ZH-EMC-890/2020\|EPI_ISL_632768 | ···················· | ···················· | ···T················· | ···················· | ···················· | ···················· |
| Q | 7147 hCoV-19/Netherlands/ZH-EMC-891/2020\|EPI_ISL_632769 | ···················· | ···················· | ····················· | ···················· | ···················· | ···················· |
| Q | 7148 hCoV-19/Netherlands/ZH-EMC-892/2020\|EPI_ISL_632770 | ···················· | ···················· | ····················· | ···················· | ···················· | ···················· |
| Q | 7149 hCoV-19/Netherlands/NB-EMC-317/2020\|EPI_ISL_632771 | ···················· | ···················· | ····················· | ···················· | ···················· | ···················· |
| Q | 7150 hCoV-19/Netherlands/NB-EMC-319/2020\|EPI_ISL_632772 | ···················· | ···················· | ····················· | ···················· | ···················· | ···················· |
| Q | 7151 hCoV-19/Netherlands/NB-EMC-320/2020\|EPI_ISL_632773 | ···················· | ···················· | ····················· | ···················· | ···················· | ···················· |

Q 7152 hCoV-19/Netherlands/NB-EMC-321/2020|EPI_ISL_632774 ···················· ···················· ···················· ···················· ···················· ····················
Q 7153 hCoV-19/Netherlands/NB-EMC-322/2020|EPI_ISL_632775 ···················· ···················· ···················· ···················· ···················· ····················
Q 7154 hCoV-19/Netherlands/un-EMC-795/2020|EPI_ISL_632776 ···················· ···················· ···················· ···················· ···················· ····················
Q 7155 hCoV-19/Netherlands/un-EMC-796/2020|EPI_ISL_632777 ···················· ···················· ···················· ···················· ···················· ····················
Q 7156 hCoV-19/Netherlands/FL-EMC-38/2020|EPI_ISL_632778| ···················· ···················· ···················· ···················· ···················· ····················
Q 7157 hCoV-19/Netherlands/UT-EMC-141/2020|EPI_ISL_632779 ···················· ···················· ···················· ···················· ···················· ····················
Q 7158 hCoV-19/Netherlands/UT-EMC-142/2020|EPI_ISL_632780 ···················· ···················· ···················· ···················· ···················· ····················
Q 7159 hCoV-19/Netherlands/FL-EMC-39/2020|EPI_ISL_632781| ···················· ···················· ···················· ···················· ···················· ····················
Q 7160 hCoV-19/Netherlands/UT-EMC-143/2020|EPI_ISL_632782 ···················· ···················· ···················· ···················· ······T············· ····················
Q 7161 hCoV-19/Netherlands/UT-EMC-144/2020|EPI_ISL_632783 ···················· ···················· ···················· ···················· ···················· ····················
Q 7162 hCoV-19/Netherlands/UT-EMC-145/2020|EPI_ISL_632784 ···················· ···················· ···················· ···················· ···················· ····················
Q 7163 hCoV-19/Netherlands/ZH-EMC-895/2020|EPI_ISL_632785 ···················· ···················· ···················· ···················· ···················· ····················
Q 7164 hCoV-19/Netherlands/ZH-EMC-896/2020|EPI_ISL_632786 ···················· ···················· ···················· ···················· ···················· ····················
Q 7165 hCoV-19/Netherlands/ZH-EMC-897/2020|EPI_ISL_632787 ···················· ···················· ···················· ···················· ···················· ····················
Q 7166 hCoV-19/Netherlands/ZH-EMC-898/2020|EPI_ISL_632788 ···················· ···················· ···················· ···················· ···················· ····················
Q 7167 hCoV-19/Netherlands/ZH-EMC-899/2020|EPI_ISL_632789 ···················· ···················· ···················· ···················· ···················· ····················
Q 7168 hCoV-19/Gibraltar/204462901/2020|EPI_ISL_632799|20 ···················· ···················· ···················· ···················· ···················· ····················
Q 7169 hCoV-19/Australia/NSW1157/2020|EPI_ISL_629010|2020 ···················· ···················· ···················· ···················· ···················· ····················
Q 7170 hCoV-19/Chile/AT-CMM-0152M/2020|EPI_ISL_629011|202 ···················· ···················· ···················· ···················· ···················· ····················
Q 7171 hCoV-19/Chile/RM-CMM-0166M/2020|EPI_ISL_629012|202 ···················· ···················· ···················· ···················· ···················· ····················
Q 7172 hCoV-19/Chile/RM-CMM-0184M/2020|EPI_ISL_629013|202 ···················· ···················· ···················· ···················· ···················· ····················
Q 7173 hCoV-19/Chile/RM-CMM-0039M/2020|EPI_ISL_629017|202 ···················· ···················· ···················· ···················· ···················· ····················
Q 7174 hCoV-19/Chile/RM-CMM-0069M/2020|EPI_ISL_629018|202 ···················· ···················· ···················· ···················· ···················· ····················
Q 7175 hCoV-19/Chile/RM-CMM-0071M/2020|EPI_ISL_629019|202 ···················· ···················· ···················· ···················· ···················· ····················
Q 7176 hCoV-19/Chile/RM-CMM-0007M/2020|EPI_ISL_629020|202 ···················· ···················· ···················· ···················· ···················· ····················
Q 7177 hCoV-19/Chile/RM-CMM-0089M/2020|EPI_ISL_629021|202 ···················· ···················· ···················· ···················· ···················· ····················
Q 7178 hCoV-19/Chile/RM-CMM-0096M/2020|EPI_ISL_629022|202 ···················· ···················· ···················· ···················· ···················· ····················
Q 7179 hCoV-19/Chile/RM-CMM-0000B/2020|EPI_ISL_629023|202 ···················· ···················· ···················· ···················· ···················· ····················
Q 7180 hCoV-19/Chile/RM-CMM-0000C/2020|EPI_ISL_629024|202 ···················· ···················· ···················· ···················· ···················· ····················
Q 7181 hCoV-19/Chile/RM-CMM-0000D/2020|EPI_ISL_629025|202 ···················· ···················· ···················· ···················· ···················· ····················
Q 7182 hCoV-19/Chile/RM-CMM-0000E/2020|EPI_ISL_629026|202 ···················· ···················· ···················· ···················· ···················· ····················
Q 7183 hCoV-19/Chile/RM-CMM-0000F/2020|EPI_ISL_629027|202 ···················· ···················· ···················· ···················· ···················· ····················
Q 7184 hCoV-19/Chile/RM-CMM-0023C/2020|EPI_ISL_629028|202 ···················· ···················· ···················· ···················· ···················· ····················
Q 7185 hCoV-19/Australia/NSW1158/2020|EPI_ISL_629029|2020 ···················· ···················· ···················· ···················· ······T············· ···············A····
Q 7186 hCoV-19/England/MILK-ACC1A9/2020|EPI_ISL_629170|20 ···················· ···················· ···················· ···················· ······T············· ····················
Q 7187 hCoV-19/England/MILK-ACBC7C/2020|EPI_ISL_629171|20 ···················· ···················· ···················· ···················· ···················· ····················
Q 7188 hCoV-19/England/MILK-ACC103/2020|EPI_ISL_629172|20 ···················· ···················· ···················· ···················· ···················· ····················
Q 7189 hCoV-19/England/MILK-ACE259/2020|EPI_ISL_629173|20 ···················· ·A·················· ···················· ···················· ···················· ···A················
Q 7190 hCoV-19/England/MILK-ACC0F7/2020|EPI_ISL_629174|20 ···················· ···················· ···················· ···················· ···················· ····················
Q 7191 hCoV-19/England/MILK-ACBE76/2020|EPI_ISL_629175|20 ···················· ···················· ···················· ···················· ···················· ····················
Q 7192 hCoV-19/England/MILK-ACC0CA/2020|EPI_ISL_629176|20 ···················· ···················· ···················· ···················· ···················· ····················
Q 7193 hCoV-19/England/MILK-ACED85/2020|EPI_ISL_629178|20 ···················· ···················· ···················· ···················· ···················· ····················
Q 7194 hCoV-19/England/MILK-ACEC88/2020|EPI_ISL_629179|20 ···················· ···················· ···················· ···················· ······T············· ····················
Q 7195 hCoV-19/England/MILK-ACC042/2020|EPI_ISL_629186|20 ···················· ···················· ···················· ···················· ······T············· ····················
Q 7196 hCoV-19/England/MILK-ACC33A/2020|EPI_ISL_629187|20 ···················· ···················· ···················· ···················· ···················· ····················
Q 7197 hCoV-19/England/MILK-AC7D6E/2020|EPI_ISL_629188|20 ···················· ···················· ···················· ···················· ······T············· ····················
Q 7198 hCoV-19/England/MILK-ACE2D1/2020|EPI_ISL_629189|20 ···················· ·A·················· ···················· ···················· ···················· ···A················
Q 7199 hCoV-19/England/MILK-ACC2C4/2020|EPI_ISL_629190|20 ···················· ···················· ···················· ···················· ···················· ····················
Q 7200 hCoV-19/England/MILK-AC92D6/2020|EPI_ISL_629191|20 ···················· ···················· ···················· ···················· ···················· ····················
Q 7201 hCoV-19/England/MILK-AC8626/2020|EPI_ISL_629192|20 ···················· ···················· ···················· ···················· ···················· ····················
Q 7202 hCoV-19/England/MILK-ACC2D3/2020|EPI_ISL_629193|20 ···················· ···················· ···················· ···················· ···················· ····················
Q 7203 hCoV-19/England/MILK-ACED76/2020|EPI_ISL_629194|20 ···················· ···················· ···················· ···················· ···················· ····················
Q 7204 hCoV-19/England/MILK-ACCB6F/2020|EPI_ISL_629195|20 ···················· ···················· ···················· ···················· ···················· ····················
Q 7205 hCoV-19/England/MILK-ACC72F/2020|EPI_ISL_629196|20 ···················· ···················· ···················· ···················· ···················· ····················
Q 7206 hCoV-19/England/MILK-ACE417/2020|EPI_ISL_629197|20 ···················· ·A·················· ···················· ···················· ···················· ···A················
Q 7207 hCoV-19/England/MILK-ACE462/2020|EPI_ISL_629198|20 ···················· ···················· ···················· ···················· ···················· ····················
Q 7208 hCoV-19/England/QEUH-AD2D9D/2020|EPI_ISL_630001|20 ···················· ···················· ···················· ···················· ···················· ····················
Q 7209 hCoV-19/England/QEUH-AD25D1/2020|EPI_ISL_630002|20 ···················· ···················· ···················· ···················· ···················· ····················
Q 7210 hCoV-19/England/QEUH-AD1833/2020|EPI_ISL_630003|20 ···················· ···················· ···················· ···················· ···················· ····················
Q 7211 hCoV-19/England/QEUH-AD2FD3/2020|EPI_ISL_630004|20 ···················· ···················· ···················· ···················· ···················· ····················
Q 7212 hCoV-19/England/QEUH-AD63A6/2020|EPI_ISL_630005|20 ···················· ···················· ···················· ···················· ···················· ····················
Q 7213 hCoV-19/England/QEUH-AD049A/2020|EPI_ISL_630006|20 ···················· ···················· ···················· ···················· ···················· ····················
Q 7214 hCoV-19/England/MILK-ACD278/2020|EPI_ISL_630007|20 ···················· ···················· ···················· ···················· ···················· ····················
Q 7215 hCoV-19/England/MILK-ACCE75/2020|EPI_ISL_630008|20 ···················· ···················· ···················· ···················· ···················· ····················
Q 7216 hCoV-19/England/MILK-AC7663/2020|EPI_ISL_630009|20 ···················· ···················· ···················· ···················· ···················· ····················
Q 7217 hCoV-19/England/MILK-AC73A8/2020|EPI_ISL_630011|20 ···················· ···················· ···················· ···················· ···················· ····················
Q 7218 hCoV-19/England/MILK-ACDF44/2020|EPI_ISL_630012|20 ···················· ···················· ···················· ···················· ···················· ····················
Q 7219 hCoV-19/England/MILK-ACCF36/2020|EPI_ISL_630013|20 ···················· ···················· ···················· ···················· ······T············· ····················
Q 7220 hCoV-19/England/MILK-ACDF17/2020|EPI_ISL_630015|20 ···················· ·A·················· ···················· ···················· ···················· ···A················
Q 7221 hCoV-19/England/MILK-ACD111/2020|EPI_ISL_630159|20 ···················· ···················· ···················· ···················· ···················· ····················
Q 7222 hCoV-19/England/MILK-AC76BE/2020|EPI_ISL_630160|20 ···················· ···················· ···················· ···················· ······T············· ····················
Q 7223 hCoV-19/England/MILK-ACCFEB/2020|EPI_ISL_630161|20 ···················· ···················· ···················· ···················· ···················· ····················
Q 7224 hCoV-19/England/MILK-ACCD1E/2020|EPI_ISL_630163|20 ···················· ···················· ···················· ···················· ···················· ····················
Q 7225 hCoV-19/England/MILK-ACDFEA/2020|EPI_ISL_630164|20 ···················· ···················· ···················· ···················· ···················· ····················
Q 7226 hCoV-19/England/MILK-AC9494/2020|EPI_ISL_630165|20 ···················· ···················· ···················· ···················· ···················· ····················
Q 7227 hCoV-19/England/MILK-ACD9FB/2020|EPI_ISL_630166|20 ···················· ···················· ···················· ···················· ······T············· ····················
Q 7228 hCoV-19/England/MILK-AC752A/2020|EPI_ISL_630168|20 ···················· ···················· ···················· ···················· ···················· ····················
Q 7229 hCoV-19/England/MILK-ACD7C4/2020|EPI_ISL_630169|20 ···················· ···················· ···················· ···················· ···················· ····················
Q 7230 hCoV-19/England/MILK-ACCF81/2020|EPI_ISL_630175|20 ···················· ···················· ···················· ···················· ···················· ····················
Q 7231 hCoV-19/England/MILK-ACCD96/2020|EPI_ISL_630180|20 ···················· ···················· ···················· ···················· ···················· ····················

| | | | | | | | |
|---|---|---|---|---|---|---|---|
| Q | 7232 hCoV-19/England/MILK-ACCF72/2020\|EPI_ISL_630181\|20 | ···················· | ···················· | ···················· | ···················· | ···················· | ···················· |
| Q | 7233 hCoV-19/England/MILK-ACD06F/2020\|EPI_ISL_630182\|20 | ···················· | ···················· | ···················· | ···················· | ······T············· | ···················· |
| Q | 7234 hCoV-19/England/MILK-ACCD69/2020\|EPI_ISL_630183\|20 | ···················· | ···················· | ···················· | ···················· | ···················· | ···················· |
| Q | 7235 hCoV-19/England/MILK-ACDF08/2020\|EPI_ISL_630184\|20 | ···················· | ···················· | ···················· | ···················· | ···················· | ···················· |
| Q | 7236 hCoV-19/England/MILK-ACCDC3/2020\|EPI_ISL_630185\|20 | ···················· | ···················· | ···················· | ···················· | ···················· | ···················· |
| Q | 7237 hCoV-19/England/MILK-AC7487/2020\|EPI_ISL_630186\|20 | ···················· | ···················· | ···················· | ···················· | ···················· | ···················· |
| Q | 7238 hCoV-19/England/MILK-ACDB8C/2020\|EPI_ISL_630188\|20 | ···················· | ···················· | ···················· | ···················· | ···················· | ···················· |
| Q | 7239 hCoV-19/England/MILK-ACDCF2/2020\|EPI_ISL_630190\|20 | ···················· | ···················· | ···················· | ···················· | ···················· | ···················· |
| Q | 7240 hCoV-19/England/MILK-ACD287/2020\|EPI_ISL_630192\|20 | ···················· | ···················· | ···················· | ···················· | ···················· | ···················· |
| Q | 7241 hCoV-19/England/MILK-AC72E7/2020\|EPI_ISL_630193\|20 | ···················· | ···················· | ···················· | ···················· | ···················· | ···················· |
| Q | 7242 hCoV-19/England/MILK-ACCE1B/2020\|EPI_ISL_630194\|20 | ···················· | ···················· | ···················· | ···················· | ···················· | ···················· |
| Q | 7243 hCoV-19/England/MILK-ACDC6B/2020\|EPI_ISL_630196\|20 | ···················· | ···················· | ···················· | ···················· | ···················· | ···················· |
| Q | 7244 hCoV-19/England/MILK-ACD18A/2020\|EPI_ISL_630197\|20 | ···················· | ···················· | ···················· | ···················· | ···················· | ···················· |
| Q | 7245 hCoV-19/England/MILK-AC751B/2020\|EPI_ISL_630198\|20 | ···················· | ···················· | ···················· | ···················· | ···················· | ···················· |
| Q | 7246 hCoV-19/England/MILK-ACD982/2020\|EPI_ISL_630199\|20 | ···················· | ·A·······A·········· | ···················· | ···················· | ···················· | ···················· |
| Q | 7247 hCoV-19/Germany/NW-HHU-256/2020\|EPI_ISL_631344\|202 | ···················· | ···················· | ···················· | ···················· | ···················· | ···················· |
| Q | 7248 hCoV-19/Germany/NW-HHU-257/2020\|EPI_ISL_631345\|202 | ···················· | ·A·················· | ···················· | ···················· | ···················· | ···················· |
| Q | 7249 hCoV-19/Germany/NW-HHU-258/2020\|EPI_ISL_631346\|202 | ···················· | ···················· | ···················· | ···················· | ···················· | ···················· |
| Q | 7257 hCoV-19/USA/WI-WSLH-200739/2020\|EPI_ISL_631495\|202 | ···················· | ···················· | ···················· | ···················· | ···················· | ···················· |
| Q | 7260 hCoV-19/USA/WI-WSLH-200743/2020\|EPI_ISL_631498\|202 | ···················· | ···················· | ···················· | ···················· | ···················· | ···················· |
| Q | 7262 hCoV-19/Netherlands/ZH-EMC-787/2020\|EPI_ISL_632656 | ···················· | ···················· | ···················· | ···················· | ···················· | ···················· |
| Q | 7263 hCoV-19/Netherlands/ZH-EMC-788/2020\|EPI_ISL_632657 | ···················· | ···················· | ···················· | ···················· | ······T············· | ···················· |
| Q | 7264 hCoV-19/Netherlands/NB-EMC-305/2020\|EPI_ISL_632658 | ···················· | ···················· | ···················· | ···················· | ···················· | ···················· |
| Q | 7265 hCoV-19/Netherlands/NB-EMC-306/2020\|EPI_ISL_632659 | ···················· | ···················· | ···················· | ···················· | ···················· | ···················· |
| Q | 7266 hCoV-19/Netherlands/NB-EMC-307/2020\|EPI_ISL_632660 | ···················· | ···················· | ···················· | ···················· | ···················· | ···················· |
| Q | 7267 hCoV-19/Netherlands/NB-EMC-308/2020\|EPI_ISL_632661 | ···················· | ···················· | ···················· | ···················· | ···················· | ···················· |
| Q | 7268 hCoV-19/Netherlands/NB-EMC-309/2020\|EPI_ISL_632662 | ···················· | ···················· | ···················· | ···················· | ···················· | ···················· |
| Q | 7269 hCoV-19/Netherlands/NB-EMC-310/2020\|EPI_ISL_632663 | ···················· | ···················· | ···················· | ···················· | ···················· | ···················· |
| Q | 7270 hCoV-19/Netherlands/NB-EMC-311/2020\|EPI_ISL_632664 | ···················· | ···················· | ···················· | ···················· | ···················· | ···················· |
| Q | 7271 hCoV-19/Netherlands/NB-EMC-312/2020\|EPI_ISL_632665 | ···················· | ····G··············· | ···················· | ···················· | ···················· | ···················· |
| Q | 7272 hCoV-19/Netherlands/NB-EMC-314/2020\|EPI_ISL_632666 | ···················· | ···················· | ···················· | ···················· | ···················· | ···················· |
| Q | 7273 hCoV-19/Netherlands/NB-EMC-315/2020\|EPI_ISL_632667 | ···················· | ···················· | ···················· | ···················· | ···················· | ···················· |
| Q | 7274 hCoV-19/Netherlands/NB-EMC-316/2020\|EPI_ISL_632668 | ···················· | ···················· | ···················· | ···················· | ···················· | ···················· |
| Q | 7275 hCoV-19/Netherlands/GE-EMC-326/2020\|EPI_ISL_632669 | ···················· | ···················· | ···················· | ···················· | ···················· | ···················· |
| Q | 7276 hCoV-19/Netherlands/ZH-EMC-789/2020\|EPI_ISL_632670 | ···················· | ···················· | ···················· | ···················· | ···················· | ···················· |
| Q | 7277 hCoV-19/Netherlands/ZH-EMC-790/2020\|EPI_ISL_632671 | ···················· | ···················· | ···················· | ···················· | ···················· | ···················· |
| Q | 7278 hCoV-19/Netherlands/ZH-EMC-791/2020\|EPI_ISL_632672 | ···················· | ···················· | ···················· | ···················· | ···················· | ···················· |
| Q | 7279 hCoV-19/Netherlands/ZH-EMC-792/2020\|EPI_ISL_632673 | ···················· | ···················· | ···················· | ···················· | ···················· | ···················· |
| Q | 7280 hCoV-19/Netherlands/ZH-EMC-793/2020\|EPI_ISL_632674 | ···················· | ···················· | ···················· | ···················· | ···················· | ···················· |
| Q | 7281 hCoV-19/Netherlands/ZH-EMC-794/2020\|EPI_ISL_632675 | ···················· | ···················· | ···················· | ···················· | ···················· | ···················· |
| Q | 7282 hCoV-19/Netherlands/ZH-EMC-797/2020\|EPI_ISL_632676 | ···················· | ···················· | ···················· | ···················· | ···················· | ···················· |
| Q | 7283 hCoV-19/Netherlands/ZH-EMC-798/2020\|EPI_ISL_632677 | ···················· | ···················· | ···················· | ···················· | ···················· | ···················· |
| Q | 7284 hCoV-19/Netherlands/ZH-EMC-828/2020\|EPI_ISL_632700 | ···················· | ···················· | ···················· | ···················· | ···················· | ···················· |
| Q | 7285 hCoV-19/Netherlands/ZH-EMC-831/2020\|EPI_ISL_632701 | ···················· | ···················· | ···················· | ···················· | ···················· | ···················· |
| Q | 7286 hCoV-19/Netherlands/ZH-EMC-832/2020\|EPI_ISL_632702 | ···················· | ···················· | ···················· | ···················· | ······T····T········ | ···················· |
| Q | 7287 hCoV-19/Netherlands/ZH-EMC-833/2020\|EPI_ISL_632703 | ···················· | ···················· | ···················· | ···················· | ···················· | ···················· |
| Q | 7288 hCoV-19/Netherlands/ZH-EMC-834/2020\|EPI_ISL_632704 | ···················· | ···················· | ···················· | ···················· | ···················· | ···················· |
| Q | 7289 hCoV-19/Netherlands/ZH-EMC-839/2020\|EPI_ISL_632705 | ···················· | ···················· | ···················· | ···················· | ······T············· | ···················· |
| Q | 7290 hCoV-19/Netherlands/ZH-EMC-841/2020\|EPI_ISL_632706 | ···················· | ···················· | ···················· | ···················· | ······T············· | ···················· |
| Q | 7291 hCoV-19/Netherlands/ZH-EMC-843/2020\|EPI_ISL_632707 | ···················· | ···················· | ···················· | ···················· | ···················· | ···················· |
| Q | 7292 hCoV-19/Netherlands/ZH-EMC-844/2020\|EPI_ISL_632708 | ···················· | ···················· | ···················· | ···················· | ···················· | ···················· |
| Q | 7293 hCoV-19/Netherlands/ZH-EMC-845/2020\|EPI_ISL_632709 | ···················· | ···················· | ···················· | ···················· | ···················· | ···················· |
| Q | 7294 hCoV-19/Netherlands/ZH-EMC-846/2020\|EPI_ISL_632710 | ···················· | ···················· | ···················· | ···················· | ···················· | ···················· |
| Q | 7295 hCoV-19/Netherlands/GE-EMC-327/2020\|EPI_ISL_632711 | ···················· | ···················· | ···················· | ···················· | ···················· | ···················· |
| Q | 7296 hCoV-19/Netherlands/GE-EMC-328/2020\|EPI_ISL_632712 | ···················· | ···················· | ···················· | ···················· | ···················· | ···················· |
| Q | 7297 hCoV-19/Netherlands/GE-EMC-329/2020\|EPI_ISL_632713 | ···················· | ···················· | ···················· | ···················· | ···················· | ···················· |
| Q | 7298 hCoV-19/Netherlands/ZH-EMC-847/2020\|EPI_ISL_632714 | ···················· | ···················· | ···················· | ···················· | ···················· | ···················· |
| Q | 7299 hCoV-19/Netherlands/LI-EMC-97/2020\|EPI_ISL_632715\| | ···················· | ···················· | ···················· | ···················· | ···················· | ···················· |
| Q | 7300 hCoV-19/Netherlands/ZH-EMC-848/2020\|EPI_ISL_632716 | ···················· | ···················· | ···················· | ···················· | ···················· | ···················· |
| Q | 7301 hCoV-19/Netherlands/ZH-EMC-849/2020\|EPI_ISL_632717 | ···················· | ···················· | ···················· | ···················· | ···················· | ···················· |
| Q | 7302 hCoV-19/Netherlands/ZH-EMC-850/2020\|EPI_ISL_632718 | ···················· | ···················· | ···················· | ···················· | ···················· | ···················· |
| Q | 7303 hCoV-19/Netherlands/ZH-EMC-851/2020\|EPI_ISL_632719 | ···················· | G··················· | ···················· | ···················· | ···················· | ···················· |
| Q | 7304 hCoV-19/Netherlands/UT-EMC-124/2020\|EPI_ISL_632720 | ···················· | ···················· | ···················· | ···················· | ···················· | ···················· |
| Q | 7305 hCoV-19/Netherlands/UT-EMC-127/2020\|EPI_ISL_632721 | ···················· | ···················· | ···················· | ···················· | ······T············· | ···················· |
| Q | 7306 hCoV-19/Netherlands/GE-EMC-338/2020\|EPI_ISL_632722 | ···················· | ···················· | ···················· | ···················· | ······T············· | ···················· |
| Q | 7307 hCoV-19/Netherlands/UT-EMC-129/2020\|EPI_ISL_632723 | ···················· | ···················· | ···················· | ···················· | ···················· | ···················· |
| Q | 7308 hCoV-19/Netherlands/UT-EMC-131/2020\|EPI_ISL_632724 | ···················· | ···················· | ···················· | ···················· | ···················· | ···················· |
| Q | 7309 hCoV-19/Netherlands/FL-EMC-32/2020\|EPI_ISL_632725\| | ···················· | ···················· | ···················· | ···················· | ···················· | ···················· |
| Q | 7310 hCoV-19/Netherlands/UT-EMC-132/2020\|EPI_ISL_632726 | ···················· | ···················· | ···················· | ···················· | ······T············· | ···················· |
| Q | 7311 hCoV-19/Netherlands/UT-EMC-133/2020\|EPI_ISL_632727 | ···················· | ···················· | ···················· | ···················· | ···················· | ···················· |
| Q | 7312 hCoV-19/Netherlands/FL-EMC-33/2020\|EPI_ISL_632728\| | ···················· | ···················· | ···················· | ···················· | ···················· | ···················· |
| Q | 7313 hCoV-19/Netherlands/FL-EMC-34/2020\|EPI_ISL_632729\| | ···················· | ···················· | ···················· | ···················· | ···················· | ···················· |
| Q | 7314 hCoV-19/Netherlands/UT-EMC-134/2020\|EPI_ISL_632730 | ···················· | ···················· | ···················· | ···················· | ···················· | ···················· |
| Q | 7315 hCoV-19/Netherlands/UT-EMC-138/2020\|EPI_ISL_632731 | ···················· | ···················· | ···················· | ···················· | ···················· | ···················· |
| Q | 7316 hCoV-19/Netherlands/UT-EMC-139/2020\|EPI_ISL_632732 | ···················· | ···················· | ···················· | ···················· | ······T············· | ···················· |
| Q | 7317 hCoV-19/England/QEUH-AD148A/2020\|EPI_ISL_633987\|20 | ···················· | ···················· | ···················· | ···················· | ···················· | ···················· |
| Q | 7318 hCoV-19/England/QEUH-AD4FB3/2020\|EPI_ISL_633989\|20 | ···················· | ·A·················· | ···················· | ···················· | ···················· | ···A················ |
| Q | 7319 hCoV-19/England/QEUH-AD327F/2020\|EPI_ISL_633990\|20 | ···················· | ···················· | ···················· | ···················· | ···················· | ···················· |
| Q | 7320 hCoV-19/England/QEUH-AD12AE/2020\|EPI_ISL_633991\|20 | ···················· | ···················· | ···················· | ···················· | ···················· | ···················· |
| Q | 7321 hCoV-19/England/QEUH-AD393E/2020\|EPI_ISL_633992\|20 | ···················· | ···················· | ···················· | ···················· | ···················· | ···················· |

Q 7322 hCoV-19/England/QEUH-AD4F0E/2020|EPI_ISL_633993|20 ···················· ···················· ····················· ···················· ···················· ····················
Q 7323 hCoV-19/England/QEUH-AD1AD3/2020|EPI_ISL_633994|20 ···················· ···················· ····················· ···················· ···················· ····················
Q 7324 hCoV-19/England/QEUH-AD4F2C/2020|EPI_ISL_633995|20 ···················· ···················· ····················· ···················· ···················· ····················
Q 7325 hCoV-19/England/QEUH-AD0E23/2020|EPI_ISL_633996|20 ···················· ···················· ····················· ···················· ···················· ····················
Q 7326 hCoV-19/England/QEUH-AD15D2/2020|EPI_ISL_633997|20 ···················· ···················· ····················· ···················· ···················· ····················
Q 7327 hCoV-19/England/QEUH-AD35FE/2020|EPI_ISL_633998|20 ···················· ···················· ····················· ···················· ···················· ····················
Q 7328 hCoV-19/England/QEUH-AD5D21/2020|EPI_ISL_633999|20 ···················· ···················· ····················· ···················· ······T············· ····················
Q 7329 hCoV-19/USA/CA-ALSR-3786/2020|EPI_ISL_636200|2020- ···················· ···················· ····················· ···················· ···················· ····················
Q 7330 hCoV-19/USA/CA-ALSR-3788/2020|EPI_ISL_636201|2020- ···················· ·A·················· ····················· ···················· ···················· ····················
Q 7331 hCoV-19/USA/CA-ALSR-3789/2020|EPI_ISL_636202|2020- ···················· ···················· ····················· ···················· ···················· ····················
Q 7332 hCoV-19/USA/CA-ALSR-3792/2020|EPI_ISL_636203|2020- ···················· ·A·················· ····················· ···················· ···················· ····················
Q 7333 hCoV-19/USA/CA-ALSR-3793/2020|EPI_ISL_636204|2020- ···················· ·A·················· ····················· ···················· ···················· ····················
Q 7334 hCoV-19/USA/CA-ALSR-3795/2020|EPI_ISL_636205|2020- ···················· ·A·················· ····················· ···················· ···················· ····················
Q 7335 hCoV-19/USA/CA-ALSR-3798/2020|EPI_ISL_636206|2020- ···················· ·A·················· ····················· ···················· ···················· ····················
Q 7336 hCoV-19/USA/CA-ALSR-3800/2020|EPI_ISL_636207|2020- ···················· ·A·················· ····················· ···················· ···················· ····················
Q 7337 hCoV-19/USA/CA-ALSR-3801/2020|EPI_ISL_636208|2020- ···················· ·A·················· ····················· ···················· ···················· ····················
Q 7338 hCoV-19/USA/CA-ALSR-3802/2020|EPI_ISL_636209|2020- ···················· ···················· ····················· ···················· ···················· ····················
Q 7339 hCoV-19/USA/CA-ALSR-3803/2020|EPI_ISL_636210|2020- ···················· ···················· ····················· ···················· ···················· ····················
Q 7340 hCoV-19/USA/CA-ALSR-3805/2020|EPI_ISL_636211|2020- ···················· ·A·················· ····················· ···················· ···················· ····················
Q 7341 hCoV-19/USA/CA-ALSR-3806/2020|EPI_ISL_636212|2020- ···················· ·A·················· ····················· ···················· ···················· ····················
Q 7342 hCoV-19/USA/CA-ALSR-3807/2020|EPI_ISL_636213|2020- ···················· ·A·················· ····················· ···················· ···················· ····················
Q 7343 hCoV-19/USA/CA-ALSR-3808/2020|EPI_ISL_636214|2020- ···················· ·A·················· ····················· ···················· ···················· ····················
Q 7344 hCoV-19/USA/CA-ALSR-3809/2020|EPI_ISL_636215|2020- ···················· ·A·················· ····················· ···················· ···················· ····················
Q 7345 hCoV-19/USA/CA-ALSR-3811/2020|EPI_ISL_636216|2020- ···················· ·A·················· ····················· ···················· ···················· ····················
Q 7346 hCoV-19/USA/CA-ALSR-3812/2020|EPI_ISL_636217|2020- ···················· ·A·················· ····················· ···················· ···················· ····················
Q 7347 hCoV-19/USA/CA-ALSR-3813/2020|EPI_ISL_636218|2020- ···················· ·A·················· ····················· ···················· ···················· ····················
Q 7348 hCoV-19/USA/CA-ALSR-3815/2020|EPI_ISL_636219|2020- ···················· ·A·················· ····················· ···················· ···················· ····················
Q 7349 hCoV-19/USA/CA-ALSR-3816/2020|EPI_ISL_636220|2020- ···················· ·A·················· ····················· ···················· ···················· ····················
Q 7350 hCoV-19/USA/CA-ALSR-3817/2020|EPI_ISL_636221|2020- ···················· ·A·················· ····················· ···················· ···················· ····················
Q 7351 hCoV-19/USA/CA-ALSR-3818/2020|EPI_ISL_636222|2020- ···················· ·A·················· ····················· ···················· ···················· ····················
Q 7352 hCoV-19/USA/CA-ALSR-3819/2020|EPI_ISL_636223|2020- ···················· ·A·················· ····················· ···················· ···················· ····················
Q 7353 hCoV-19/USA/CA-ALSR-3820/2020|EPI_ISL_636224|2020- ···················· ···················· ····················· ···················· ···················· ····················
Q 7354 hCoV-19/USA/CA-ALSR-3821/2020|EPI_ISL_636225|2020- ···················· ·A·················· ····················· ···················· ···················· ····················
Q 7355 hCoV-19/USA/CA-ALSR-3822/2020|EPI_ISL_636226|2020- ···················· ·A·················· ····················· ···················· ···················· ····················
Q 7356 hCoV-19/USA/CA-ALSR-3823/2020|EPI_ISL_636227|2020- ···················· ···················· ····················· ···················· ···················· ····················
Q 7357 hCoV-19/USA/CA-ALSR-3824/2020|EPI_ISL_636228|2020- ···················· ·A·················· ····················· ···················· ···················· ····················
Q 7358 hCoV-19/USA/CA-ALSR-3827/2020|EPI_ISL_636229|2020- ···················· ···················· ····················· ···················· ···················· ····················
Q 7359 hCoV-19/USA/CA-ALSR-3828/2020|EPI_ISL_636230|2020- ···················· ·A·················· ····················· ···················· ···················· ····················
Q 7360 hCoV-19/USA/CA-ALSR-3831/2020|EPI_ISL_636231|2020- ···················· ·A·················· ····················· ···················· ···················· ····················
Q 7361 hCoV-19/USA/CA-ALSR-3832/2020|EPI_ISL_636232|2020- ···················· ·A··A··············· ····················· ···················· ···················· ····················
Q 7362 hCoV-19/USA/CA-ALSR-3834/2020|EPI_ISL_636233|2020- ···················· ·A·················· ····················· ···················· ···················· ····················
Q 7363 hCoV-19/USA/CA-ALSR-3835/2020|EPI_ISL_636234|2020- ···················· ···················· ····················· ···················· ···················· ····················
Q 7364 hCoV-19/USA/CA-ALSR-3838/2020|EPI_ISL_636235|2020- ···················· ···················· ····················· ···················· ···················· ····················
Q 7365 hCoV-19/USA/CA-ALSR-3840/2020|EPI_ISL_636236|2020- ···················· ···················· ····················· ···················· ···················· ····················
Q 7366 hCoV-19/USA/CA-ALSR-3841/2020|EPI_ISL_636237|2020- ···················· ·A·················· ····················· ···················· ···················· ····················
Q 7367 hCoV-19/USA/CA-ALSR-3844/2020|EPI_ISL_636239|2020- ···················· ·A·················· ····················· ···················· ···················· ····················
Q 7368 hCoV-19/USA/CA-ALSR-3847/2020|EPI_ISL_636240|2020- ···················· ·A·················· ····················· ···················· ···················· ····················
Q 7369 hCoV-19/USA/CA-ALSR-3848/2020|EPI_ISL_636241|2020- ···················· ·A·················· ····················· ···················· ···················· ····················
Q 7370 hCoV-19/USA/CA-ALSR-3849/2020|EPI_ISL_636242|2020- ···················· ·A·················· ····················· ···················· ···················· ····················
Q 7371 hCoV-19/USA/CA-ALSR-3850/2020|EPI_ISL_636243|2020- ···················· ·A·················· ····················· ···················· ···················· ····················
Q 7372 hCoV-19/USA/CA-ALSR-3851/2020|EPI_ISL_636244|2020- ···················· ·A·················· ····················· ···················· ···················· ····················
Q 7373 hCoV-19/USA/CA-ALSR-3855/2020|EPI_ISL_636245|2020- ···················· ···················· ····················· ···················· ···················· ····················
Q 7374 hCoV-19/USA/CA-ALSR-3856/2020|EPI_ISL_636246|2020- ···················· ·A·················· ····················· ···················· ···················· ····················
Q 7375 hCoV-19/USA/CA-ALSR-3857/2020|EPI_ISL_636247|2020- ···················· ·A·················· ····················· ···················· ···················· ····················
Q 7376 hCoV-19/USA/CA-ALSR-3861/2020|EPI_ISL_636248|2020- ···················· ·A·················· ····················· ···················· ···················· ····················
Q 7377 hCoV-19/USA/CA-ALSR-3863/2020|EPI_ISL_636250|2020- ···················· ·A·················· ····················· ···················· ···················· ····················
Q 7378 hCoV-19/USA/CA-ALSR-3868/2020|EPI_ISL_636251|2020- ···················· ·A·················· ····················· ···················· ···················· ····················
Q 7379 hCoV-19/USA/CA-ALSR-3870/2020|EPI_ISL_636252|2020- ···················· ·A·················· ····················· ···················· ···················· ····················
Q 7380 hCoV-19/USA/CA-ALSR-3872/2020|EPI_ISL_636253|2020- ···················· ·A·················· ····················· ···················· ···················· ····················
Q 7381 hCoV-19/USA/CA-ALSR-3873/2020|EPI_ISL_636254|2020- ···················· ·A·················· ····················· ···················· ···················· ····················
Q 7382 hCoV-19/USA/CA-ALSR-3875/2020|EPI_ISL_636255|2020- ···················· ···················· ····················· ···················· ···················· ····················
Q 7383 hCoV-19/USA/CA-ALSR-3877/2020|EPI_ISL_636256|2020- ···················· ···················· ····················· ···················· ···················· ····················
Q 7384 hCoV-19/USA/CA-ALSR-3880/2020|EPI_ISL_636258|2020- ···················· ···················· ····················· ···················· ···················· ····················
Q 7385 hCoV-19/USA/CA-ALSR-3885/2020|EPI_ISL_636260|2020- ···················· ·A·················· ····················· ···················· ···················· ····················
Q 7386 hCoV-19/USA/CA-ALSR-3886/2020|EPI_ISL_636261|2020- ···················· ·A·················· ····················· ···················· ···················· ····················
Q 7387 hCoV-19/USA/CA-ALSR-3887/2020|EPI_ISL_636262|2020- ···················· ···················· ····················· ···················· ···················· ····················
Q 7388 hCoV-19/USA/CA-ALSR-3891/2020|EPI_ISL_636263|2020- ···················· ···················· ····················· ···················· ···················· ····················
Q 7389 hCoV-19/USA/CA-ALSR-3895/2020|EPI_ISL_636264|2020- ···················· ·A·················· ····················· ···················· ···················· ····················
Q 7390 hCoV-19/USA/CA-ALSR-3898/2020|EPI_ISL_636265|2020- ···················· ···················· ····················· ···················· ···················· ····················
Q 7391 hCoV-19/USA/CA-ALSR-3899/2020|EPI_ISL_636266|2020- ···················· ·A·················· ····················· ···················· ···················· ····················
Q 7392 hCoV-19/Jordan/AM-ALSR-3901/2020|EPI_ISL_636267|20 ···················· ···················· ····················· ···················· ···················· ····················
Q 7393 hCoV-19/Jordan/IR-ALSR-3902/2020|EPI_ISL_636268|20 ···················· ·A·················· ····················· ···················· ···················· ··A·················
Q 7394 hCoV-19/Jordan/AM-ALSR-3903/2020|EPI_ISL_636269|20 ···················· ···················· ····················· ···················· ···················· ····················
Q 7395 hCoV-19/Jordan/AM-ALSR-3904/2020|EPI_ISL_636270|20 ···················· ···················· ····················· ···················· ···················· ····················
Q 7396 hCoV-19/Jordan/AM-ALSR-3905/2020|EPI_ISL_636271|20 ···················· ···················· ····················· ···················· ···················· ····················
Q 7397 hCoV-19/Jordan/AM-ALSR-3906/2020|EPI_ISL_636272|20 ···················· ···················· ····················· ···················· ···················· ····················
Q 7398 hCoV-19/Jordan/AM-ALSR-3907/2020|EPI_ISL_636273|20 ···················· ···················· ····················· ···················· ···················· ····················
Q 7399 hCoV-19/Jordan/AM-ALSR-3908/2020|EPI_ISL_636274|20 ···················· ·A·················· ····················· ···················· ···················· ··A·················
Q 7400 hCoV-19/Jordan/AM-ALSR-3909/2020|EPI_ISL_636275|20 ···················· ···················· ····················· ···················· ···················· ····················
Q 7401 hCoV-19/Jordan/AM-ALSR-3910/2020|EPI_ISL_636276|20 ···················· ···················· ····················· ···················· ···················· ····················

| | | | | | | | |
|---|---|---|---|---|---|---|---|
| Q | 7402 hCoV-19/Jordan/AM-ALSR-3911/2020\|EPI_ISL_636277\|20 | ···················· | ···················· | ····················· | ···················· | ···················· | ···················· |
| Q | 7403 hCoV-19/Jordan/AM-ALSR-3912/2020\|EPI_ISL_636278\|20 | ···················· | ·A·················· | ····················· | ···················· | ···················· | ··A················· |
| Q | 7404 hCoV-19/Jordan/AM-ALSR-3913/2020\|EPI_ISL_636279\|20 | ···················· | ···················· | ····················· | ···················· | ···················· | ···················· |
| Q | 7405 hCoV-19/Jordan/AM-ALSR-3914/2020\|EPI_ISL_636280\|20 | ···················· | ···················· | ····················· | ···················· | ···················· | ···················· |
| Q | 7406 hCoV-19/Jordan/AM-ALSR-3915/2020\|EPI_ISL_636281\|20 | ···················· | ···················· | ····················· | ···················· | ···················· | ···················· |
| Q | 7407 hCoV-19/Jordan/AM-ALSR-3916/2020\|EPI_ISL_636282\|20 | ···················· | ···················· | ····················· | ···················· | ···················· | ···················· |
| Q | 7408 hCoV-19/Jordan/IR-ALSR-3917/2020\|EPI_ISL_636283\|20 | ···················· | ·A·················· | ····················· | ···················· | ···················· | ··A················· |
| Q | 7409 hCoV-19/Jordan/AM-ALSR-3918/2020\|EPI_ISL_636284\|20 | ···················· | ·A·················· | ····················· | ···················· | ···················· | ··A················· |
| Q | 7410 hCoV-19/Jordan/AM-ALSR-3919/2020\|EPI_ISL_636285\|20 | ···················· | ·A·················· | ····················· | ···················· | ···················· | ··A················· |
| Q | 7411 hCoV-19/Jordan/AM-ALSR-3920/2020\|EPI_ISL_636286\|20 | ···················· | ···················· | ····················· | ···················· | ···················· | ···················· |
| Q | 7412 hCoV-19/Jordan/AM-ALSR-3921/2020\|EPI_ISL_636287\|20 | ···················· | ···················· | ····················· | ···················· | ···················· | ···················· |
| Q | 7413 hCoV-19/Jordan/AM-ALSR-3922/2020\|EPI_ISL_636288\|20 | ···················· | ···················· | ····················· | ···················· | ···················· | ···················· |
| Q | 7414 hCoV-19/Jordan/AM-ALSR-3923/2020\|EPI_ISL_636289\|20 | ···················· | ···················· | ····················· | ···················· | ···················· | ···················· |
| Q | 7415 hCoV-19/Jordan/AM-ALSR-3925/2020\|EPI_ISL_636290\|20 | ···················· | ···················· | ····················· | ···················· | ···················· | ···················· |
| Q | 7416 hCoV-19/Jordan/AM-ALSR-3926/2020\|EPI_ISL_636291\|20 | ···················· | ·A·················· | ····················· | ···················· | ···················· | ··A················· |
| Q | 7417 hCoV-19/Jordan/AM-ALSR-3927/2020\|EPI_ISL_636292\|20 | ···················· | ···················· | ····················· | ···················· | ···················· | ···················· |
| Q | 7418 hCoV-19/Jordan/AM-ALSR-3928/2020\|EPI_ISL_636293\|20 | ···················· | ···················· | ····················· | ···················· | ···················· | ···················· |
| Q | 7419 hCoV-19/Jordan/AM-ALSR-3929/2020\|EPI_ISL_636294\|20 | ···················· | ···················· | ····················· | ···················· | ···················· | ···················· |
| Q | 7420 hCoV-19/Jordan/AM-ALSR-3930/2020\|EPI_ISL_636295\|20 | ···················· | ···················· | ····················· | ···················· | ···················· | ···················· |
| Q | 7421 hCoV-19/Jordan/AM-ALSR-3931/2020\|EPI_ISL_636296\|20 | ···················· | ···················· | ····················· | ···················· | ···················· | ···················· |
| Q | 7422 hCoV-19/Jordan/AM-ALSR-3932/2020\|EPI_ISL_636297\|20 | ···················· | ···················· | ····················· | ···················· | ···················· | ···················· |
| Q | 7423 hCoV-19/Jordan/AM-ALSR-3933/2020\|EPI_ISL_636298\|20 | ···················· | ···················· | ····················· | ···················· | ···················· | ···················· |
| Q | 7424 hCoV-19/Jordan/AM-ALSR-3935/2020\|EPI_ISL_636299\|20 | ···················· | ···················· | ····················· | ···················· | ···················· | ···················· |
| Q | 7425 hCoV-19/Australia/NSW1148/2020\|EPI_ISL_629001\|2020 | ···················· | ·A·················· | ····················· | ···················· | ···················· | ··A················· |
| Q | 7426 hCoV-19/Australia/NSW1149/2020\|EPI_ISL_629002\|2020 | ···················· | ···················· | ····················· | ···················· | ···················· | ···················· |
| Q | 7427 hCoV-19/Australia/NSW1150/2020\|EPI_ISL_629003\|2020 | ···················· | ···················· | ····················· | ···················· | ···················· | ···················· |
| Q | 7428 hCoV-19/Australia/NSW1151/2020\|EPI_ISL_629004\|2020 | ···················· | ···················· | ····················· | ···················· | ···················· | ···················· |
| Q | 7429 hCoV-19/Australia/NSW1152/2020\|EPI_ISL_629005\|2020 | ···················· | ···················· | ····················· | ···················· | ···················· | ···················· |
| Q | 7430 hCoV-19/Australia/NSW1153/2020\|EPI_ISL_629006\|2020 | ···················· | ···················· | ····················· | ···················· | ···················· | ···················· |
| Q | 7431 hCoV-19/Australia/NSW1154/2020\|EPI_ISL_629007\|2020 | ···················· | ···················· | ····················· | ···················· | ···················· | ···················· |
| Q | 7432 hCoV-19/Australia/NSW1155/2020\|EPI_ISL_629008\|2020 | ···················· | ···················· | ····················· | ···················· | ···················· | ···················· |
| Q | 7433 hCoV-19/Australia/NSW1156/2020\|EPI_ISL_629009\|2020 | ···················· | ···················· | ····················· | ···················· | ···················· | ···················· |
| Q | 7434 hCoV-19/Chile/RM-CMM-0194M/2020\|EPI_ISL_629014\|202 | ···················· | ···················· | ····················· | ···················· | ···················· | ···················· |
| Q | 7435 hCoV-19/Chile/RM-CMM-0257M/2020\|EPI_ISL_629015\|202 | ···················· | ···················· | ····················· | ···················· | ···················· | ···················· |
| Q | 7436 hCoV-19/Chile/RM-CMM-0263M/2020\|EPI_ISL_629016\|202 | ···················· | ···················· | ····················· | ···················· | ···················· | ···················· |
| Q | 7437 hCoV-19/Australia/NSW1159/2020\|EPI_ISL_629030\|2020 | ···················· | ···················· | ····················· | ···················· | ···················· | ···················· |
| Q | 7438 hCoV-19/Russia/OMS-ORINFI-4761V-10K/2020\|EPI_ISL_6 | ···················· | ···················· | ····················· | ···················· | ···················· | ···················· |
| Q | 7439 hCoV-19/Russia/OMS-ORINFI-4761V-11K/2020\|EPI_ISL_6 | ···················· | ···················· | ····················· | ···················· | ···················· | ···················· |
| Q | 7440 hCoV-19/Russia/OMS-ORINFI-4768V-12K/2020\|EPI_ISL_6 | ···················· | ···················· | ····················· | ···················· | ···················· | ···················· |
| Q | 7441 hCoV-19/Russia/OMS-ORINFI-4761V/2020\|EPI_ISL_62903 | ···················· | ···················· | ····················· | ···················· | ···················· | ···················· |
| Q | 7442 hCoV-19/USA/MD-MDH-0255/2020\|EPI_ISL_629036\|2020-1 | ···················· | ···················· | ····················· | ···················· | ···················· | ···················· |
| Q | 7443 hCoV-19/USA/MD-MDH-0256/2020\|EPI_ISL_629037\|2020-1 | ···················· | ···················· | ····················· | ···················· | ···················· | ···················· |
| Q | 7444 hCoV-19/USA/MD-MDH-0257/2020\|EPI_ISL_629038\|2020-1 | ···················· | ···················· | ····················· | ···················· | ···················· | ···················· |
| Q | 7445 hCoV-19/USA/MD-MDH-0258/2020\|EPI_ISL_629039\|2020-1 | ···················· | ···················· | ····················· | ···················· | ···················· | ···················· |
| Q | 7446 hCoV-19/USA/MD-MDH-0259/2020\|EPI_ISL_629040\|2020-1 | ···················· | ············A······· | ····················· | ···················· | ···················· | ···················· |
| Q | 7447 hCoV-19/USA/MD-MDH-0260/2020\|EPI_ISL_629041\|2020-1 | ···················· | ·A·················· | ····················· | ···················· | ···················· | ···················· |
| Q | 7448 hCoV-19/USA/MD-MDH-0261/2020\|EPI_ISL_629042\|2020-1 | ···················· | ·A·················· | ····················· | ···················· | ···················· | ···················· |
| Q | 7449 hCoV-19/USA/MD-MDH-0262/2020\|EPI_ISL_629043\|2020-1 | ···················· | ·A·················· | ····················· | ···················· | ···················· | ···················· |
| Q | 7450 hCoV-19/USA/MD-MDH-0263/2020\|EPI_ISL_629044\|2020-1 | ···················· | ·A·················· | ····················· | ···················· | ···················· | ···················· |
| Q | 7451 hCoV-19/USA/MD-MDH-0264/2020\|EPI_ISL_629045\|2020-1 | ···················· | ·A·················· | ····················· | ···················· | ···················· | ···················· |
| Q | 7452 hCoV-19/USA/MD-MDH-0265/2020\|EPI_ISL_629046\|2020-1 | ···················· | ·A·················· | ····················· | ···················· | ···················· | ···················· |
| Q | 7453 hCoV-19/USA/MD-MDH-0266/2020\|EPI_ISL_629047\|2020-1 | ···················· | ·A·················· | ····················· | ···················· | ···················· | ···················· |
| Q | 7454 hCoV-19/USA/MD-MDH-0267/2020\|EPI_ISL_629048\|2020-1 | ···················· | ·A·················· | ····················· | ···················· | ···················· | ···················· |
| Q | 7455 hCoV-19/USA/MD-MDH-0268/2020\|EPI_ISL_629049\|2020-1 | ···················· | ·A·················· | ····················· | ···················· | ···················· | ···················· |
| Q | 7456 hCoV-19/USA/MD-MDH-0280/2020\|EPI_ISL_629060\|2020-1 | ···················· | ···················· | ····················· | ···················· | ···················· | ···················· |
| Q | 7457 hCoV-19/USA/MD-MDH-0281/2020\|EPI_ISL_629061\|2020-1 | ···················· | ···················· | ····················· | ···················· | ···················· | ···················· |
| Q | 7458 hCoV-19/USA/MD-MDH-0282/2020\|EPI_ISL_629062\|2020-1 | ···················· | ···················· | ····················· | ···················· | ···················· | ···················· |
| Q | 7459 hCoV-19/USA/MD-MDH-0283/2020\|EPI_ISL_629063\|2020-1 | ···················· | ·A·················· | ····················· | ···················· | ···················· | ···················· |
| Q | 7460 hCoV-19/USA/MD-MDH-0284/2020\|EPI_ISL_629064\|2020-1 | ···················· | ···················· | ····················· | ···················· | ···················· | ···················· |
| Q | 7461 hCoV-19/USA/MD-MDH-0286/2020\|EPI_ISL_629066\|2020-1 | ···················· | ···················· | ····················· | ···················· | ···················· | ···················· |
| Q | 7462 hCoV-19/USA/MD-MDH-0287/2020\|EPI_ISL_629067\|2020-1 | ···················· | ···················· | ····················· | ···················· | ···················· | ···················· |
| Q | 7463 hCoV-19/USA/MD-MDH-0288/2020\|EPI_ISL_629068\|2020-1 | ···················· | ···················· | ····················· | ···················· | ···················· | ···················· |
| Q | 7464 hCoV-19/France/ARA-SC024/2020\|EPI_ISL_629090\|2020- | ···················· | ···················· | ····················· | ···················· | ···················· | ···················· |
| Q | 7465 hCoV-19/France/ARA-SC027/2020\|EPI_ISL_629091\|2020- | ···················· | ···················· | ····················· | ···················· | ···················· | ···················· |
| Q | 7466 hCoV-19/France/ARA-SC033/2020\|EPI_ISL_629092\|2020- | ···················· | ···················· | ····················· | ···················· | ···················· | ···················· |
| Q | 7467 hCoV-19/France/ARA-SC035/2020\|EPI_ISL_629093\|2020- | ···················· | ···················· | ····················· | ···················· | ···················· | ···················· |
| Q | 7468 hCoV-19/France/ARA-SC036/2020\|EPI_ISL_629094\|2020- | ···················· | ···················· | ····················· | ···················· | ···················· | ···················· |
| Q | 7469 hCoV-19/France/ARA-SC040/2020\|EPI_ISL_629095\|2020- | ···················· | ···················· | ····················· | ···················· | ···················· | ···················· |
| Q | 7470 hCoV-19/France/ARA-SC046/2020\|EPI_ISL_629096\|2020- | ···················· | ···················· | ····················· | ···················· | ···················· | ···················· |
| Q | 7471 hCoV-19/France/ARA-SC050/2020\|EPI_ISL_629097\|2020- | ···················· | ···················· | ····················· | ···················· | ···················· | ···················· |
| Q | 7472 hCoV-19/France/ARA-SC053/2020\|EPI_ISL_629098\|2020- | ···················· | ···················· | ····················· | ···················· | ···················· | ···················· |
| Q | 7473 hCoV-19/France/ARA-SC054/2020\|EPI_ISL_629099\|2020- | ···················· | ···················· | ····················· | ···················· | ···················· | ···················· |
| Q | 7474 hCoV-19/England/MILK-AC8301/2020\|EPI_ISL_630016\|20 | ···················· | ···················· | ····················· | ···················· | ···················· | ···················· |
| Q | 7475 hCoV-19/England/MILK-AC738A/2020\|EPI_ISL_630017\|20 | ···················· | ···················· | ····················· | ···················· | ···················· | ···················· |
| Q | 7476 hCoV-19/England/MILK-ACD0C9/2020\|EPI_ISL_630018\|20 | ···················· | ···················· | ····················· | ···················· | ···················· | ···················· |
| Q | 7477 hCoV-19/England/MILK-ACCE2A/2020\|EPI_ISL_630020\|20 | ···················· | ···················· | ····················· | ···················· | ···················· | ···················· |
| Q | 7478 hCoV-19/England/MILK-ACCD2D/2020\|EPI_ISL_630021\|20 | ···················· | ···················· | ····················· | ···················· | ···················· | ···················· |
| Q | 7479 hCoV-19/England/MILK-ACDCD4/2020\|EPI_ISL_630023\|20 | ···················· | ···················· | ····················· | ···················· | ···················· | ···················· |
| Q | 7480 hCoV-19/England/MILK-ACDC4D/2020\|EPI_ISL_630024\|20 | ···················· | ···················· | ····················· | ···················· | ···················· | ···················· |
| Q | 7481 hCoV-19/England/MILK-ACCFFA/2020\|EPI_ISL_630025\|20 | ···················· | ···················· | ····················· | ···················· | ···················· | ···················· |

Q 7482 hCoV-19/England/MILK-AC74B4/2020|EPI_ISL_630027|20 ···················· ···················· ······················ ···················· ···················· ····················
Q 7483 hCoV-19/England/MILK-AC9449/2020|EPI_ISL_630028|20 ···················· ···················· ······················ ···················· ···················· ····················
Q 7484 hCoV-19/England/MILK-ACD2D2/2020|EPI_ISL_630029|20 ···················· ···················· ······················ ···················· ···················· ····················
Q 7485 hCoV-19/England/MILK-AC7469/2020|EPI_ISL_630030|20 ···················· ···················· ······················ ···················· ···················· ····················
Q 7486 hCoV-19/England/MILK-ACD937/2020|EPI_ISL_630031|20 ···················· ···················· ······················ ···················· ···················· ····················
Q 7487 hCoV-19/England/MILK-ACDEDE/2020|EPI_ISL_630032|20 ···················· ···················· ······················ ···················· ···················· ····················
Q 7488 hCoV-19/England/MILK-ACCFBE/2020|EPI_ISL_630033|20 ···················· ···················· ······················ ···················· ···················· ····················
Q 7489 hCoV-19/England/MILK-ACDE56/2020|EPI_ISL_630034|20 ···················· ···················· ······················ ···················· ···················· ····················
Q 7490 hCoV-19/England/MILK-ACDE0B/2020|EPI_ISL_630035|20 ···················· ···················· ······················ ···················· ······T············· ····················
Q 7491 hCoV-19/England/MILK-AC93F1/2020|EPI_ISL_630049|20 ···················· ·A·················· ······················ ···················· ···················· ···A················
Q 7492 hCoV-19/England/MILK-ACDC7A/2020|EPI_ISL_630050|20 ···················· ···················· ······················ ···················· ···················· ····················
Q 7493 hCoV-19/England/MILK-AC9379/2020|EPI_ISL_630052|20 ···················· ·A·················· ······················ ···················· ···················· ····················
Q 7494 hCoV-19/England/MILK-ACCCA8/2020|EPI_ISL_630053|20 ···················· ···················· ······················ ···················· ······T············· ····················
Q 7495 hCoV-19/England/MILK-ACD13F/2020|EPI_ISL_630054|20 ···················· ···················· ······················ ···················· ···················· ····················
Q 7496 hCoV-19/England/MILK-AC743C/2020|EPI_ISL_630055|20 ···················· ·A·················· ······················ ···················· ···················· ····················
Q 7497 hCoV-19/England/MILK-AC9476/2020|EPI_ISL_630056|20 ···················· ···················· ······················ ···················· ···················· ····················
Q 7498 hCoV-19/England/MILK-ACDC10/2020|EPI_ISL_630057|20 ···················· ···················· ······················ ···················· ···················· ····················
Q 7499 hCoV-19/England/MILK-ACDE1A/2020|EPI_ISL_630058|20 ···················· ···················· ······················ ···················· ···················· ····················
Q 7500 hCoV-19/England/MILK-ACCD0F/2020|EPI_ISL_630059|20 ···················· ···················· ······················ ···················· ···················· ····················
Q 7501 hCoV-19/England/MILK-ACDF9F/2020|EPI_ISL_630080|20 ···················· ···················· ······················ ···················· ···················· ····················
Q 7502 hCoV-19/England/MILK-ACD797/2020|EPI_ISL_630081|20 ···················· ···················· ······················ ···················· ······T············· ····················
Q 7503 hCoV-19/England/MILK-ACDD0E/2020|EPI_ISL_630082|20 ···················· ···················· ······················ ···················· ···················· ····················
Q 7504 hCoV-19/England/MILK-ACD102/2020|EPI_ISL_630083|20 ···················· ·A·················· ······················ ···················· ···················· ···A················
Q 7505 hCoV-19/England/MILK-AC7654/2020|EPI_ISL_630084|20 ···················· ···················· ······················ ···················· ···················· ····················
Q 7506 hCoV-19/England/MILK-ACD621/2020|EPI_ISL_630085|20 ···················· ···················· ······················ ···················· ···················· ····················
Q 7507 hCoV-19/England/MILK-ACDDA4/2020|EPI_ISL_630087|20 ···················· ···················· ······················ ···················· ······T············· ····················
Q 7508 hCoV-19/England/MILK-AC76FA/2020|EPI_ISL_630088|20 ···················· ···················· ······················ ···················· ···················· ····················
Q 7509 hCoV-19/England/MILK-ACE013/2020|EPI_ISL_630089|20 ···················· ···················· ······················ ···················· ···················· ····················
Q 7510 hCoV-19/England/MILK-AC9467/2020|EPI_ISL_630090|20 ···················· ···················· ······················ ···················· ···················· ····················
Q 7511 hCoV-19/England/MILK-ACE13E/2020|EPI_ISL_630092|20 ···················· ···················· ······················ ···················· ···················· ····················
Q 7512 hCoV-19/England/MILK-AC73F3/2020|EPI_ISL_630093|20 ···················· ···················· ······················ ···················· ······T············· ····················
Q 7513 hCoV-19/England/MILK-ACE004/2020|EPI_ISL_630094|20 ···················· ···················· ······················ ···················· ···················· ····················
Q 7514 hCoV-19/England/MILK-ACDCB6/2020|EPI_ISL_630096|20 ···················· ···················· ······················ ···················· ···················· ····················
Q 7515 hCoV-19/England/MILK-ACDBC8/2020|EPI_ISL_630097|20 ···················· ···················· ······················ ···················· ···················· ····················
Q 7516 hCoV-19/England/MILK-ACDD3B/2020|EPI_ISL_630098|20 ···················· ···················· ······················ ···················· ···················· ····················
Q 7517 hCoV-19/England/MILK-ACE1A7/2020|EPI_ISL_630099|20 ···················· ···················· ······················ ···················· ···················· ····················
Q 7518 hCoV-19/Germany/NW-UBI-46/2020|EPI_ISL_631300|2020 ···················· ···················· ······················ ···················· ···················· ····················
Q 7519 hCoV-19/Germany/NW-UBI-48/2020|EPI_ISL_631301|2020 ···················· ···················· ······················ ···················· ···················· ····················
Q 7520 hCoV-19/Germany/NW-UBI-49/2020|EPI_ISL_631302|2020 ···················· ···················· ······················ ···················· ···················· ····················
Q 7521 hCoV-19/Germany/NW-UBI-50/2020|EPI_ISL_631303|2020 ···················· ···················· ······················ ···················· ···················· ····················
Q 7522 hCoV-19/Germany/NW-HHU-301/2020|EPI_ISL_631304|202 ···················· ···················· ······················ ···················· ···················· ····················
Q 7523 hCoV-19/Germany/NW-HHU-302/2020|EPI_ISL_631305|202 ···················· ···················· ······················ ···················· ···················· ···············A····
Q 7524 hCoV-19/Germany/NW-HHU-303/2020|EPI_ISL_631306|202 ···················· ···················· ······················ ···················· ···················· ···············A····
Q 7525 hCoV-19/Germany/NW-HHU-304/2020|EPI_ISL_631307|202 ···················· ···················· ······················ ···················· ···················· ····················
Q 7526 hCoV-19/Germany/NW-HHU-305/2020|EPI_ISL_631308|202 ···················· ···················· ······················ ···················· ···················· ···············A····
Q 7527 hCoV-19/Germany/NW-HHU-306/2020|EPI_ISL_631309|202 ···················· ···················· ······················ ···················· ···················· ···············A····
Q 7528 hCoV-19/Germany/NW-HHU-307/2020|EPI_ISL_631310|202 ···················· ···················· ······················ ···················· ···················· ····················
Q 7529 hCoV-19/Germany/NW-HHU-308/2020|EPI_ISL_631311|202 ···················· ···················· ······················ ···················· ···················· ···············A····
Q 7530 hCoV-19/Germany/NW-HHU-309/2020|EPI_ISL_631312|202 ···················· ···················· ······················ ···················· ···················· ····················
Q 7531 hCoV-19/Germany/NW-HHU-310/2020|EPI_ISL_631313|202 ···················· ···················· ······················ ···················· ···················· ····················
Q 7532 hCoV-19/Germany/NW-HHU-311/2020|EPI_ISL_631314|202 ···················· ···················· ······················ ···················· ···················· ····················
Q 7533 hCoV-19/Germany/NW-HHU-312/2020|EPI_ISL_631315|202 ···················· ···················· ······················ ···················· ···················· ····················
Q 7534 hCoV-19/Germany/NW-HHU-313/2020|EPI_ISL_631316|202 ···················· ···················· ······················ ···················· ···················· ····················
Q 7535 hCoV-19/Germany/NW-HHU-314/2020|EPI_ISL_631317|202 ···················· ···················· ······················ ···················· ······T············· ····················
Q 7536 hCoV-19/Germany/NW-HHU-315/2020|EPI_ISL_631318|202 ···················· ···················· ······················ ···················· ···················· ····················
Q 7537 hCoV-19/Germany/NW-HHU-316/2020|EPI_ISL_631319|202 ···················· ···················· ······················ ···················· ···················· ····················
Q 7538 hCoV-19/Germany/NW-HHU-320/2020|EPI_ISL_631320|202 ···················· ···················· ······················ ···················· ···················· ····················
Q 7539 hCoV-19/Germany/NW-HHU-277/2020|EPI_ISL_631321|202 ···················· ···················· ······················ ···················· ···················· ···············A····
Q 7540 hCoV-19/Germany/NW-HHU-278/2020|EPI_ISL_631322|202 ···················· ···················· ······················ ···················· ···················· ····················
Q 7541 hCoV-19/Germany/NW-HHU-280/2020|EPI_ISL_631324|202 ···················· ···················· ······················ ···················· ···················· ····················
Q 7542 hCoV-19/Germany/NW-HHU-260/2020|EPI_ISL_631348|202 ···················· ···················· ······················ ···················· ···················· ····················
Q 7543 hCoV-19/Germany/NW-HHU-261/2020|EPI_ISL_631349|202 ···················· ···················· ······················ ···················· ···················· ····················
Q 7544 hCoV-19/Germany/NW-HHU-262/2020|EPI_ISL_631350|202 ···················· ···················· ······················ ···················· ···················· ····················
Q 7545 hCoV-19/Germany/NW-HHU-263/2020|EPI_ISL_631351|202 ···················· ···················· ······················ ···················· ···················· ····················
Q 7546 hCoV-19/Germany/NW-HHU-264/2020|EPI_ISL_631352|202 ···················· ···················· ······················ ···················· ···················· ····················
Q 7547 hCoV-19/Germany/NW-HHU-265/2020|EPI_ISL_631353|202 ···················· ···················· ······················ ···················· ···················· ····················
Q 7548 hCoV-19/Germany/NW-HHU-266/2020|EPI_ISL_631354|202 ···················· ···················· ······················ ···················· ···················· ····················
Q 7549 hCoV-19/Germany/NW-HHU-267/2020|EPI_ISL_631355|202 ···················· ···················· ······················ ···················· ···················· ····················
Q 7550 hCoV-19/Germany/NW-HHU-268/2020|EPI_ISL_631356|202 ···················· ···················· ······················ ···················· ···················· ····················
Q 7551 hCoV-19/Germany/NW-HHU-269/2020|EPI_ISL_631357|202 ···················· ···················· ······················ ···················· ···················· ····················
Q 7552 hCoV-19/Germany/NW-HHU-270/2020|EPI_ISL_631358|202 ···················· ···················· ······················ ···················· ···················· ····················
Q 7553 hCoV-19/Germany/NW-HHU-271/2020|EPI_ISL_631359|202 ···················· ···················· ······················ ···················· ···················· ····················
Q 7554 hCoV-19/Germany/NW-HHU-272/2020|EPI_ISL_631360|202 ···················· ·A·················· ······················ ···················· ···················· ····················
Q 7555 hCoV-19/Germany/NW-HHU-273/2020|EPI_ISL_631361|202 ···················· ···················· ······················ ···················· ···················· ····················
Q 7556 hCoV-19/Germany/NW-HHU-274/2020|EPI_ISL_631362|202 ···················· ···················· ······················ ···················· ···················· ····················
Q 7557 hCoV-19/Germany/NW-HHU-275/2020|EPI_ISL_631363|202 ···················· ···················· ······················ ···················· ···················· ····················
Q 7558 hCoV-19/Germany/NW-HHU-276/2020|EPI_ISL_631364|202 ···················· ···A················ ······················ ···················· ···················· ····················
Q 7559 hCoV-19/Germany/NW-HHU-232/2020|EPI_ISL_631365|202 ···················· ···················· ······················ ···················· ······T············· ····················
Q 7560 hCoV-19/Germany/NW-HHU-233/2020|EPI_ISL_631366|202 ···················· ···················· ······················ ···················· ······T············· ····················
Q 7561 hCoV-19/Germany/NW-HHU-234/2020|EPI_ISL_631367|202 ···················· ···················· ······················ ···················· ······T············· ····················

| | | | | | | | |
|---|---|---|---|---|---|---|---|
| Q | 7562 hCoV-19/Germany/NW-HHU-235/2020\|EPI_ISL_631368\|202 | .................... | .................... | .................... | .................... | ......T............. | .................... |
| Q | 7563 hCoV-19/Germany/NW-HHU-247/2020\|EPI_ISL_631380\|202 | .................... | ......A............. | .................... | .................... | .................... | .................... |
| Q | 7564 hCoV-19/Germany/NW-HHU-248/2020\|EPI_ISL_631381\|202 | .................... | .................... | .................... | .................... | .................... | .................... |
| Q | 7565 hCoV-19/Germany/NW-HHU-249/2020\|EPI_ISL_631382\|202 | .................... | ......A............. | .................... | .................... | .................... | .................... |
| Q | 7566 hCoV-19/Germany/NW-HHU-250/2020\|EPI_ISL_631383\|202 | .................... | ......A............. | .................... | .................... | .................... | .................... |
| Q | 7567 hCoV-19/Germany/NW-HHU-251/2020\|EPI_ISL_631384\|202 | .................... | ......A............. | .................... | .................... | .................... | .................... |
| Q | 7568 hCoV-19/Germany/NW-HHU-252/2020\|EPI_ISL_631385\|202 | .................... | .................... | .................... | .................... | .................... | .................... |
| Q | 7569 hCoV-19/Germany/NW-HHU-253/2020\|EPI_ISL_631386\|202 | .................... | .................... | .................... | .................... | .................... | .................... |
| Q | 7570 hCoV-19/Germany/NW-HHU-254/2020\|EPI_ISL_631387\|202 | .................... | .................... | .................... | .................... | .................... | .................... |
| Q | 7571 hCoV-19/Germany/NW-HHU-255/2020\|EPI_ISL_631388\|202 | .................... | .................... | .................... | .................... | ......T............. | .................... |
| Q | 7573 hCoV-19/Netherlands/ZH-EMC-775/2020\|EPI_ISL_632612 | .................... | .................... | .................... | .................... | .................... | .................... |
| Q | 7574 hCoV-19/Netherlands/ZH-EMC-776/2020\|EPI_ISL_632613 | .................... | .................... | .................... | .................... | .................... | .................... |
| Q | 7575 hCoV-19/Netherlands/ZH-EMC-777/2020\|EPI_ISL_632614 | .................... | ......A............. | .................... | .................... | .................... | .................... |
| Q | 7576 hCoV-19/Netherlands/ZH-EMC-778/2020\|EPI_ISL_632615 | .................... | ......A............. | .................... | .................... | .................... | .................... |
| Q | 7577 hCoV-19/Netherlands/ZH-EMC-779/2020\|EPI_ISL_632616 | .................... | .................... | .................... | .................... | .................... | .................... |
| Q | 7578 hCoV-19/Netherlands/ZH-EMC-780/2020\|EPI_ISL_632617 | .................... | .................... | .................... | .................... | .................... | .................... |
| Q | 7579 hCoV-19/Netherlands/ZH-EMC-781/2020\|EPI_ISL_632618 | .................... | .................... | .................... | .................... | .................... | .................... |
| Q | 7580 hCoV-19/Netherlands/ZH-EMC-782/2020\|EPI_ISL_632619 | .................... | .................... | .................... | .................... | .................... | .................... |
| Q | 7581 hCoV-19/Netherlands/ZH-EMC-783/2020\|EPI_ISL_632620 | .................... | .................... | .................... | .................... | .................... | .................... |
| Q | 7582 hCoV-19/Netherlands/ZH-EMC-784/2020\|EPI_ISL_632621 | .................... | .................... | .................... | .................... | .................... | .................... |
| Q | 7583 hCoV-19/Netherlands/ZH-EMC-785/2020\|EPI_ISL_632622 | .................... | .................... | .................... | .................... | .................... | .................... |
| Q | 7584 hCoV-19/Netherlands/UT-EMC-107/2020\|EPI_ISL_632623 | .................... | .................... | .................... | .................... | .................... | .................... |
| Q | 7585 hCoV-19/Netherlands/GE-EMC-307/2020\|EPI_ISL_632624 | .................... | .................... | .................... | .................... | .................... | .................... |
| Q | 7586 hCoV-19/Netherlands/UT-EMC-108/2020\|EPI_ISL_632625 | .................... | .................... | .................... | .................... | .................... | .................... |
| Q | 7587 hCoV-19/Netherlands/UT-EMC-147/2020\|EPI_ISL_632626 | .................... | .................... | .................... | .................... | .................... | .................... |
| Q | 7588 hCoV-19/Netherlands/GE-EMC-308/2020\|EPI_ISL_632627 | .................... | .................... | .................... | .................... | .................... | .................... |
| Q | 7589 hCoV-19/Netherlands/UT-EMC-109/2020\|EPI_ISL_632628 | .................... | .................... | .................... | .................... | ......T............. | .................... |
| Q | 7590 hCoV-19/Netherlands/UT-EMC-110/2020\|EPI_ISL_632629 | .................... | .................... | .................... | .................... | .................... | .................... |
| Q | 7591 hCoV-19/Netherlands/UT-EMC-111/2020\|EPI_ISL_632630 | .................... | .................... | .................... | .................... | .................... | .................... |
| Q | 7592 hCoV-19/Netherlands/GE-EMC-309/2020\|EPI_ISL_632631 | .................... | .................... | .................... | .................... | .................... | .................... |
| Q | 7593 hCoV-19/Netherlands/FL-EMC-29/2020\|EPI_ISL_632632\| | .................... | .................... | .................... | .................... | .................... | .................... |
| Q | 7594 hCoV-19/Netherlands/GE-EMC-310/2020\|EPI_ISL_632633 | .................... | .................... | .................... | .................... | .................... | .................... |
| Q | 7595 hCoV-19/Netherlands/UT-EMC-112/2020\|EPI_ISL_632634 | .................... | .................... | .................... | .................... | .................... | .................... |
| Q | 7596 hCoV-19/Netherlands/UT-EMC-113/2020\|EPI_ISL_632635 | .................... | .................... | .................... | .................... | .................... | .................... |
| Q | 7597 hCoV-19/Netherlands/GE-EMC-312/2020\|EPI_ISL_632636 | .................... | .................... | .................... | .................... | .................... | .................... |
| Q | 7598 hCoV-19/Netherlands/UT-EMC-114/2020\|EPI_ISL_632637 | .................... | .................... | .................... | .................... | .................... | .................... |
| Q | 7599 hCoV-19/Netherlands/UT-EMC-115/2020\|EPI_ISL_632638 | .................... | .................... | .................... | .................... | .................... | .................... |
| Q | 7600 hCoV-19/Netherlands/GE-EMC-313/2020\|EPI_ISL_632639 | .................... | .................... | .................... | .................... | .................... | .................... |
| Q | 7601 hCoV-19/Netherlands/UT-EMC-117/2020\|EPI_ISL_632640 | .................... | .A....A............. | .................... | .................... | .................... | .................... |
| Q | 7602 hCoV-19/Netherlands/UT-EMC-118/2020\|EPI_ISL_632641 | .................... | .................... | .................... | .................... | .................... | .................... |
| Q | 7603 hCoV-19/Netherlands/GE-EMC-349/2020\|EPI_ISL_632642 | .................... | .................... | .................... | .................... | .................... | .................... |
| Q | 7604 hCoV-19/Netherlands/GE-EMC-314/2020\|EPI_ISL_632643 | .................... | .................... | .................... | .................... | .................... | .................... |
| Q | 7605 hCoV-19/Netherlands/UT-EMC-119/2020\|EPI_ISL_632644 | .................... | .A.................. | .................... | .................... | .................... | ...A................ |
| Q | 7606 hCoV-19/Netherlands/UT-EMC-120/2020\|EPI_ISL_632645 | .................... | .................... | .................... | .................... | ......T............. | .................... |
| Q | 7607 hCoV-19/Netherlands/GE-EMC-315/2020\|EPI_ISL_632646 | .................... | .................... | .................... | .................... | .................... | .................... |
| Q | 7608 hCoV-19/Netherlands/GE-EMC-316/2020\|EPI_ISL_632647 | .................... | .................... | .................... | .................... | .................... | .................... |
| Q | 7609 hCoV-19/Netherlands/UT-EMC-121/2020\|EPI_ISL_632648 | .................... | .................... | .................... | .................... | .................... | .................... |
| Q | 7610 hCoV-19/Netherlands/GE-EMC-317/2020\|EPI_ISL_632649 | .................... | .................... | .................... | .................... | .................... | .................... |
| Q | 7611 hCoV-19/Netherlands/GE-EMC-318/2020\|EPI_ISL_632650 | .................... | .................... | .................... | .................... | .................... | .................... |
| Q | 7612 hCoV-19/Netherlands/GE-EMC-320/2020\|EPI_ISL_632651 | .................... | .................... | .................... | .................... | .................... | .................... |
| Q | 7613 hCoV-19/Netherlands/FL-EMC-30/2020\|EPI_ISL_632652\| | .................... | .................... | .................... | .................... | .................... | .................... |
| Q | 7614 hCoV-19/Netherlands/GE-EMC-321/2020\|EPI_ISL_632653 | .................... | .A.................. | .................... | .................... | .................... | ...A................ |
| Q | 7615 hCoV-19/Netherlands/UT-EMC-122/2020\|EPI_ISL_632654 | .................... | .................... | .................... | .................... | .................... | .................... |
| Q | 7616 hCoV-19/Netherlands/GE-EMC-323/2020\|EPI_ISL_632655 | .................... | .................... | .................... | .................... | .................... | .................... |
| Q | 7617 hCoV-19/Netherlands/ZH-EMC-799/2020\|EPI_ISL_632678 | .................... | .................... | .................... | .................... | .................... | .................... |
| Q | 7618 hCoV-19/Netherlands/ZH-EMC-800/2020\|EPI_ISL_632679 | .................... | .................... | .................... | .................... | .................... | .................... |
| Q | 7619 hCoV-19/Netherlands/ZH-EMC-801/2020\|EPI_ISL_632680 | .................... | .................... | .................... | .................... | .................... | .................... |
| Q | 7620 hCoV-19/Netherlands/ZH-EMC-802/2020\|EPI_ISL_632681 | .................... | .................... | .................... | .................... | .................... | .................... |
| Q | 7621 hCoV-19/Netherlands/ZH-EMC-803/2020\|EPI_ISL_632682 | .................... | .................... | .................... | .................... | .................... | .................... |
| Q | 7622 hCoV-19/Netherlands/ZH-EMC-804/2020\|EPI_ISL_632683 | .................... | .................... | .................... | .................... | .................... | .................... |
| Q | 7623 hCoV-19/Netherlands/ZH-EMC-805/2020\|EPI_ISL_632684 | .................... | .................... | .................... | .................... | .................... | .................... |
| Q | 7624 hCoV-19/Netherlands/ZH-EMC-806/2020\|EPI_ISL_632685 | .................... | .................... | .................... | .................... | ......T............. | .................... |
| Q | 7625 hCoV-19/Netherlands/ZH-EMC-807/2020\|EPI_ISL_632686 | .................... | .................... | .................... | .................... | .................... | .................... |
| Q | 7626 hCoV-19/Netherlands/ZH-EMC-808/2020\|EPI_ISL_632687 | .................... | .................... | .................... | .................... | .................... | .................... |
| Q | 7627 hCoV-19/Netherlands/ZH-EMC-809/2020\|EPI_ISL_632688 | .................... | .................... | .................... | .................... | .................... | .................... |
| Q | 7628 hCoV-19/Netherlands/ZH-EMC-812/2020\|EPI_ISL_632689 | .................... | .................... | .................... | .................... | .................... | .................... |
| Q | 7629 hCoV-19/Netherlands/ZH-EMC-813/2020\|EPI_ISL_632690 | .................... | .................... | .................... | .................... | .................... | .................... |
| Q | 7630 hCoV-19/Netherlands/ZH-EMC-814/2020\|EPI_ISL_632691 | .................... | .................... | .................... | .................... | .................... | .................... |
| Q | 7631 hCoV-19/Netherlands/ZH-EMC-815/2020\|EPI_ISL_632692 | .................... | .................... | .................... | .................... | .................... | .................... |
| Q | 7632 hCoV-19/Netherlands/ZH-EMC-817/2020\|EPI_ISL_632693 | .................... | .................... | .................... | .................... | .................... | .................... |
| Q | 7633 hCoV-19/Netherlands/ZH-EMC-818/2020\|EPI_ISL_632694 | .................... | .................... | .................... | .................... | .................... | .................... |
| Q | 7634 hCoV-19/Netherlands/ZH-EMC-820/2020\|EPI_ISL_632695 | .................... | .A.................. | .................... | .................... | .................... | ...A................ |
| Q | 7635 hCoV-19/Netherlands/ZH-EMC-822/2020\|EPI_ISL_632696 | .................... | .................... | .................... | .................... | .................... | .................... |
| Q | 7636 hCoV-19/Netherlands/ZH-EMC-823/2020\|EPI_ISL_632697 | .................... | .................... | .................... | .................... | .................... | .................... |
| Q | 7637 hCoV-19/Netherlands/ZH-EMC-826/2020\|EPI_ISL_632698 | .................... | .................... | .................... | .................... | .................... | .................... |
| Q | 7638 hCoV-19/Netherlands/ZH-EMC-827/2020\|EPI_ISL_632699 | .................... | .................... | .................... | .................... | .................... | .................... |
| Q | 7639 hCoV-19/England/QEUH-AD11B0/2020\|EPI_ISL_633944\|20 | .................... | .................... | .................... | .................... | .................... | .................... |
| Q | 7640 hCoV-19/England/QEUH-AD31DC/2020\|EPI_ISL_633945\|20 | .................... | .................... | .................... | .................... | .................... | .................... |
| Q | 7641 hCoV-19/England/QEUH-AD1271/2020\|EPI_ISL_633946\|20 | .................... | .................... | .................... | .................... | .................... | .................... |
| Q | 7642 hCoV-19/England/QEUH-AD08E9/2020\|EPI_ISL_633947\|20 | .................... | .................... | .................... | .................... | .................... | .................... |

| | | | | | | | |
|---|---|---|---|---|---|---|---|
| Q | 7643 hCoV-19/England/QEUH-AD1578/2020\|EPI_ISL_633948\|20 | ···················· | ···················· | ···················· | ···················· | ···················· | ···················· |
| Q | 7644 hCoV-19/England/QEUH-AD0BD1/2020\|EPI_ISL_633949\|20 | ···················· | ···················· | ···················· | ···················· | ···················· | ···················· |
| Q | 7645 hCoV-19/England/QEUH-AD13F6/2020\|EPI_ISL_633950\|20 | ···················· | ···················· | ···················· | ···················· | ···················· | ···················· |
| Q | 7646 hCoV-19/England/QEUH-AD5037/2020\|EPI_ISL_633951\|20 | ···················· | ···················· | ···················· | ···················· | ···················· | ···················· |
| Q | 7647 hCoV-19/England/QEUH-AD551A/2020\|EPI_ISL_633952\|20 | ···················· | ···················· | ···················· | ···················· | ···················· | ···················· |
| Q | 7648 hCoV-19/England/QEUH-AD0BC2/2020\|EPI_ISL_633953\|20 | ···················· | ···················· | ···················· | ···················· | ···················· | ···················· |
| Q | 7649 hCoV-19/England/QEUH-AD1253/2020\|EPI_ISL_633954\|20 | ···················· | ···················· | ···················· | ···················· | ···················· | ···················· |
| Q | 7650 hCoV-19/England/QEUH-AD58F3/2020\|EPI_ISL_633955\|20 | ···················· | ···················· | ···················· | ···················· | ···················· | ···················· |
| Q | 7651 hCoV-19/England/QEUH-AD5969/2020\|EPI_ISL_633956\|20 | ···················· | ···················· | ···················· | ···················· | ···················· | ···················· |
| Q | 7652 hCoV-19/England/QEUH-AD1332/2020\|EPI_ISL_633957\|20 | ···················· | ···················· | ···················· | ···················· | ···················· | ···················· |
| Q | 7653 hCoV-19/England/QEUH-AD3118/2020\|EPI_ISL_633958\|20 | ···················· | ···················· | ···················· | ···················· | ···················· | ···················· |
| Q | 7654 hCoV-19/England/QEUH-AD110B/2020\|EPI_ISL_633959\|20 | ···················· | ···················· | ···················· | ···················· | ···················· | ···················· |
| Q | 7655 hCoV-19/England/QEUH-AD1323/2020\|EPI_ISL_633960\|20 | ···················· | ···················· | ···················· | ···················· | ···················· | ···················· |
| Q | 7656 hCoV-19/England/QEUH-AD1341/2020\|EPI_ISL_633961\|20 | ···················· | ···················· | ···················· | ···················· | ···················· | ···················· |
| Q | 7657 hCoV-19/England/QEUH-AD0E05/2020\|EPI_ISL_633962\|20 | ···················· | ···················· | ···················· | ···················· | ···················· | ···················· |
| Q | 7658 hCoV-19/England/QEUH-AD11CF/2020\|EPI_ISL_633963\|20 | ····G··············· | ···················· | ···················· | ···················· | ···················· | ···················· |
| Q | 7659 hCoV-19/England/QEUH-AD360A/2020\|EPI_ISL_633964\|20 | ···················· | ···················· | ···················· | ···················· | ···················· | ···················· |
| Q | 7660 hCoV-19/England/QEUH-AD0B1D/2020\|EPI_ISL_633965\|20 | ···················· | ···················· | ···················· | ···················· | ···················· | ···················· |
| Q | 7661 hCoV-19/England/QEUH-AD0AA7/2020\|EPI_ISL_633966\|20 | ···················· | ···················· | ···················· | ···················· | ···················· | ···················· |
| Q | 7662 hCoV-19/England/QEUH-AD11DE/2020\|EPI_ISL_633967\|20 | ···················· | ···················· | ···················· | ···················· | ···················· | ···················· |
| Q | 7663 hCoV-19/England/QEUH-AD5019/2020\|EPI_ISL_633968\|20 | ···················· | ···················· | ···················· | ···················· | ···················· | ···················· |
| Q | 7664 hCoV-19/England/QEUH-AD38B9/2020\|EPI_ISL_633969\|20 | ···················· | ···················· | ···················· | ···················· | ···················· | ···················· |
| Q | 7665 hCoV-19/England/QEUH-AD592D/2020\|EPI_ISL_633970\|20 | ···················· | ···················· | ···················· | ···················· | ···················· | ···················· |
| Q | 7666 hCoV-19/England/QEUH-AD4E98/2020\|EPI_ISL_633971\|20 | ···················· | ···················· | ···················· | ···················· | ···················· | ···················· |
| Q | 7667 hCoV-19/England/QEUH-AD0E14/2020\|EPI_ISL_633973\|20 | ···················· | ···················· | ···················· | ···················· | ···················· | ···················· |
| Q | 7668 hCoV-19/England/QEUH-AD28C9/2020\|EPI_ISL_633974\|20 | ···················· | ···················· | ···················· | ···················· | ···················· | ···················· |
| Q | 7669 hCoV-19/England/QEUH-AD54E0/2020\|EPI_ISL_633975\|20 | ···················· | ···················· | ···················· | ···················· | ···················· | ···················· |
| Q | 7670 hCoV-19/England/QEUH-AD27EA/2020\|EPI_ISL_633976\|20 | ···················· | ···················· | ···················· | ···················· | ···················· | ···················· |
| Q | 7671 hCoV-19/England/QEUH-AD5DC7/2020\|EPI_ISL_633977\|20 | ···················· | ···················· | ···················· | ···················· | ···················· | ···················· |
| Q | 7672 hCoV-19/England/QEUH-AD5495/2020\|EPI_ISL_633978\|20 | ···················· | ···················· | ···················· | ···················· | ···················· | ···················· |
| Q | 7673 hCoV-19/England/QEUH-AD0807/2020\|EPI_ISL_633979\|20 | ···················· | ···················· | ···················· | ···················· | ···················· | ···················· |
| Q | 7674 hCoV-19/England/QEUH-AD1165/2020\|EPI_ISL_633980\|20 | ···················· | ···················· | ···················· | ···················· | ···················· | ···················· |
| Q | 7675 hCoV-19/England/QEUH-AD0D9F/2020\|EPI_ISL_633981\|20 | ···················· | ···················· | ···················· | ···················· | ···················· | ···················· |
| Q | 7676 hCoV-19/England/QEUH-AD1192/2020\|EPI_ISL_633982\|20 | ···················· | ···················· | ···················· | ···················· | ···················· | ···················· |
| Q | 7677 hCoV-19/England/QEUH-AD12CC/2020\|EPI_ISL_633983\|20 | ···················· | ···················· | ···················· | ···················· | ···················· | ···················· |
| Q | 7678 hCoV-19/England/QEUH-AD3093/2020\|EPI_ISL_633984\|20 | ···················· | ···················· | ···················· | ···················· | ···················· | ···················· |
| Q | 7679 hCoV-19/England/QEUH-AD5D4F/2020\|EPI_ISL_633985\|20 | ···················· | ···················· | ···················· | ···················· | ···················· | ···················· |
| Q | 7680 hCoV-19/England/QEUH-AD388C/2020\|EPI_ISL_633986\|20 | ···················· | ···················· | ···················· | ···················· | ···················· | ···················· |
| Q | 7681 hCoV-19/USA/MD-MDH-0269/2020\|EPI_ISL_629050\|2020-1 | ···················· | ·A·················· | ···················· | ···················· | ···················· | ···················· |
| Q | 7682 hCoV-19/USA/MD-MDH-0270/2020\|EPI_ISL_629051\|2020-1 | ···················· | ·A·················· | ···················· | ···················· | ···················· | ···················· |
| Q | 7683 hCoV-19/USA/MD-MDH-0271/2020\|EPI_ISL_629052\|2020-1 | ···················· | ·A·················· | ···················· | ···················· | ···················· | ···················· |
| Q | 7684 hCoV-19/USA/MD-MDH-0272/2020\|EPI_ISL_629053\|2020-1 | ···················· | ·A·················· | ···················· | ···················· | ···················· | ···················· |
| Q | 7685 hCoV-19/USA/MD-MDH-0273/2020\|EPI_ISL_629054\|2020-1 | ···················· | ·A·················· | ···················· | ···················· | ···················· | ···················· |
| Q | 7686 hCoV-19/USA/MD-MDH-0274/2020\|EPI_ISL_629055\|2020-1 | ···················· | ·A·················· | ···················· | ···················· | ···················· | ···················· |
| Q | 7687 hCoV-19/USA/MD-MDH-0275/2020\|EPI_ISL_629056\|2020-1 | ···················· | ·A·················· | ···················· | ···················· | ···················· | ···················· |
| Q | 7688 hCoV-19/USA/MD-MDH-0276/2020\|EPI_ISL_629057\|2020-1 | ···················· | ·A·················· | ···················· | ···················· | ···················· | ···················· |
| Q | 7689 hCoV-19/USA/MD-MDH-0277/2020\|EPI_ISL_629058\|2020-1 | ···················· | ·A·················· | ···················· | ···················· | ···················· | ···················· |
| Q | 7690 hCoV-19/USA/MD-MDH-0278/2020\|EPI_ISL_629059\|2020-1 | ···················· | ·A·················· | ···················· | ···················· | ···················· | ···················· |
| Q | 7691 hCoV-19/USA/MD-MDH-0285/2020\|EPI_ISL_629065\|2020-1 | ···················· | ···················· | ···················· | ···················· | ···················· | ···················· |
| Q | 7692 hCoV-19/USA/MD-MDH-0289/2020\|EPI_ISL_629069\|2020-1 | ···················· | ············C······· | ···················· | ···················· | ···················· | ···················· |
| Q | 7693 hCoV-19/USA/MD-MDH-0290/2020\|EPI_ISL_629070\|2020-1 | ···················· | ·A·················· | ···················· | ···················· | ···················· | ···················· |
| Q | 7694 hCoV-19/USA/MD-MDH-0291/2020\|EPI_ISL_629071\|2020-1 | ···················· | ·A·················· | ···················· | ···················· | ···················· | ···················· |
| Q | 7695 hCoV-19/USA/MD-MDH-0292/2020\|EPI_ISL_629072\|2020-1 | ···················· | ·A·················· | ···················· | ···················· | ···················· | ···················· |
| Q | 7696 hCoV-19/USA/MD-MDH-0293/2020\|EPI_ISL_629073\|2020-1 | ···················· | ·A·················· | ···················· | ···················· | ···················· | ···················· |
| Q | 7697 hCoV-19/USA/MD-MDH-0294/2020\|EPI_ISL_629074\|2020-1 | ···················· | ·A·················· | ···················· | ···················· | ···················· | ···················· |
| Q | 7698 hCoV-19/USA/MD-MDH-0295/2020\|EPI_ISL_629075\|2020-1 | ···················· | ···················· | ···················· | ···················· | ···················· | ···················· |
| Q | 7699 hCoV-19/USA/MD-MDH-0296/2020\|EPI_ISL_629076\|2020-1 | ···················· | ···················· | ···················· | ···················· | ···················· | ···················· |
| Q | 7700 hCoV-19/USA/MD-MDH-0297/2020\|EPI_ISL_629077\|2020-1 | ···················· | ···················· | ···················· | ···················· | ···················· | ···················· |
| Q | 7701 hCoV-19/USA/MD-MDH-0298/2020\|EPI_ISL_629078\|2020-1 | ···················· | ···················· | ···················· | ···················· | ···················· | ···················· |
| Q | 7702 hCoV-19/USA/MD-MDH-0299/2020\|EPI_ISL_629079\|2020-1 | ···················· | ···················· | ···················· | ···················· | ···················· | ···················· |
| Q | 7703 hCoV-19/USA/MD-MDH-0300/2020\|EPI_ISL_629080\|2020-1 | ···················· | ···················· | ···················· | ···················· | ···················· | ···················· |
| Q | 7704 hCoV-19/France/ARA-SC001/2020\|EPI_ISL_629081\|2020- | ···················· | ···················· | ···················· | ···················· | ···················· | ···················· |
| Q | 7705 hCoV-19/France/ARA-SC002/2020\|EPI_ISL_629082\|2020- | ···················· | ···················· | ···················· | ···················· | ···················· | ···················· |
| Q | 7706 hCoV-19/France/ARA-SC003/2020\|EPI_ISL_629083\|2020- | ···················· | ···················· | ···················· | ···················· | ···················· | ···················· |
| Q | 7707 hCoV-19/France/ARA-SC004/2020\|EPI_ISL_629084\|2020- | ···················· | ···················· | ···················· | ···················· | ···················· | ···················· |
| Q | 7708 hCoV-19/France/ARA-SC005/2020\|EPI_ISL_629085\|2020- | ···················· | ···················· | ···················· | ···················· | ···················· | ···················· |
| Q | 7709 hCoV-19/France/ARA-SC014/2020\|EPI_ISL_629086\|2020- | ···················· | ···················· | ···················· | ···················· | ···················· | ···················· |
| Q | 7710 hCoV-19/France/ARA-SC020/2020\|EPI_ISL_629088\|2020- | ···················· | ···················· | ···················· | ···················· | ···················· | ···················· |
| Q | 7711 hCoV-19/France/ARA-SC022/2020\|EPI_ISL_629089\|2020- | ···················· | ···················· | ···················· | ···················· | ···················· | ···················· |
| Q | 7712 hCoV-19/England/MILK-AC741E/2020\|EPI_ISL_630038\|20 | ···················· | ···················· | ···················· | ···················· | ···················· | ···················· |
| Q | 7713 hCoV-19/England/MILK-ACCE84/2020\|EPI_ISL_630039\|20 | ···················· | ···················· | ···················· | ···················· | ·····T·············· | ···················· |
| Q | 7714 hCoV-19/England/MILK-ACD7A6/2020\|EPI_ISL_630040\|20 | ···················· | ···················· | ···················· | ···················· | ···················· | ···················· |
| Q | 7715 hCoV-19/England/MILK-ACCDD2/2020\|EPI_ISL_630041\|20 | ···················· | ···················· | ···················· | ···················· | ···················· | ···················· |
| Q | 7716 hCoV-19/England/MILK-ACCF27/2020\|EPI_ISL_630042\|20 | ···················· | ···················· | ···················· | ···················· | ···················· | ···················· |
| Q | 7717 hCoV-19/England/MILK-ACD894/2020\|EPI_ISL_630043\|20 | ···················· | ···················· | ···················· | ···················· | ···················· | ···················· |
| Q | 7718 hCoV-19/England/MILK-AC737B/2020\|EPI_ISL_630044\|20 | ···················· | ···················· | ···················· | ···················· | ·····T·············· | ···················· |
| Q | 7719 hCoV-19/England/MILK-ACCF45/2020\|EPI_ISL_630045\|20 | ···················· | ···················· | ···················· | ···················· | ···················· | ···················· |
| Q | 7720 hCoV-19/England/MILK-ACDA43/2020\|EPI_ISL_630046\|20 | ···················· | ···················· | ···················· | ···················· | ···T················ | ···················· |
| Q | 7721 hCoV-19/England/MILK-ACD955/2020\|EPI_ISL_630047\|20 | ···················· | ···················· | ···················· | ···················· | ···················· | ···················· |
| Q | 7722 hCoV-19/England/MILK-ACD885/2020\|EPI_ISL_630060\|20 | ···················· | ···················· | ···················· | ···················· | ···················· | ···················· |

Q 7723 hCoV-19/England/MILK-AC75B1/2020|EPI_ISL_630061|20 ···················· ···················· ···················· ···················· ···················· ····················
Q 7724 hCoV-19/England/MILK-ACDA25/2020|EPI_ISL_630062|20 ···················· ···················· ···················· ···················· ···················· ····················
Q 7725 hCoV-19/England/MILK-ACE06E/2020|EPI_ISL_630063|20 ···················· ···················· ···················· ···················· ···················· ····················
Q 7726 hCoV-19/England/MILK-AC74A5/2020|EPI_ISL_630064|20 ···················· ···················· ···················· ···················· ···················· ····················
Q 7727 hCoV-19/England/MILK-ACE022/2020|EPI_ISL_630065|20 ···················· ·A·················· ···················· ···················· ···················· ···A················
Q 7728 hCoV-19/England/MILK-ACE110/2020|EPI_ISL_630066|20 ···················· ···················· ···················· ···················· ···················· ····················
Q 7729 hCoV-19/England/MILK-ACE0B9/2020|EPI_ISL_630067|20 ···················· ···················· ···················· ···················· ···················· ····················
Q 7730 hCoV-19/England/MILK-ACD005/2020|EPI_ISL_630068|20 ···················· ···················· ···················· ···················· ···················· ····················
Q 7731 hCoV-19/England/MILK-AC7618/2020|EPI_ISL_630069|20 ···················· ···················· ···················· ···················· ···················· ····················
Q 7732 hCoV-19/England/MILK-ACDFF9/2020|EPI_ISL_630070|20 ···················· ···················· ···················· ···················· ···················· ····················
Q 7733 hCoV-19/England/MILK-ACDADA/2020|EPI_ISL_630071|20 ···················· ···················· ···················· ···················· ···················· ····················
Q 7734 hCoV-19/England/MILK-ACDB9B/2020|EPI_ISL_630072|20 ···················· ···················· ···················· ···················· ···················· ····················
Q 7735 hCoV-19/England/MILK-ACCB23/2020|EPI_ISL_630073|20 ···················· ·A·················· ···················· ···················· ···················· ···A················
Q 7736 hCoV-19/England/MILK-ACDABC/2020|EPI_ISL_630074|20 ···················· ···················· ···················· ···················· ···················· ····················
Q 7737 hCoV-19/England/MILK-AC73D5/2020|EPI_ISL_630075|20 ···················· ···················· ···················· ···················· ···················· ····················
Q 7738 hCoV-19/England/MILK-ACDD68/2020|EPI_ISL_630076|20 ···················· ·A·················· ···················· ···················· ···················· ···A················
Q 7739 hCoV-19/England/MILK-ACD0F6/2020|EPI_ISL_630077|20 ···················· ···················· ···················· ···················· ···················· ····················
Q 7740 hCoV-19/England/MILK-ACDE29/2020|EPI_ISL_630078|20 ···················· ···················· ···················· ···················· ···················· ····················
Q 7741 hCoV-19/England/MILK-ACDA70/2020|EPI_ISL_630079|20 ···················· ···················· ···················· ···················· ···················· ····················
Q 7742 hCoV-19/England/MILK-ABBF49/2020|EPI_ISL_631211|20 ···················· ···················· ···················· ···················· ···················· ····················
Q 7743 hCoV-19/England/ALDP-ABD2C6/2020|EPI_ISL_631212|20 ···················· ···················· ···················· ···················· ···················· ····················
Q 7744 hCoV-19/England/MILK-ABBE3D/2020|EPI_ISL_631214|20 ···················· ···················· ···················· ···················· ···················· ····················
Q 7745 hCoV-19/England/ALDP-AC5D42/2020|EPI_ISL_631215|20 ···················· ···················· ···················· ···················· ···················· ····················
Q 7746 hCoV-19/England/MILK-ABC2B8/2020|EPI_ISL_631216|20 ···················· ···················· ···················· ···················· ······T············· ····················
Q 7747 hCoV-19/England/ALDP-AC5DBB/2020|EPI_ISL_631217|20 ···················· ···················· ···················· ···················· ···················· ····················
Q 7748 hCoV-19/England/MILK-ABC29A/2020|EPI_ISL_631218|20 ···················· ···················· ···················· ···················· ···················· ····················
Q 7749 hCoV-19/England/CAMC-AAF7F5/2020|EPI_ISL_631219|20 ···················· ···················· ···················· ···················· ···················· ····················
Q 7750 hCoV-19/England/CAMC-AAFF84/2020|EPI_ISL_631220|20 ···················· ···················· ···················· ···················· ···················· ····················
Q 7751 hCoV-19/England/CAMC-AAF977/2020|EPI_ISL_631221|20 ···················· ···················· ···················· ···················· ···················· ····················
Q 7752 hCoV-19/England/ALDP-ABD26C/2020|EPI_ISL_631222|20 ···················· ···················· ···················· ···················· ···················· ····················
Q 7753 hCoV-19/England/MILK-ABBCCA/2020|EPI_ISL_631223|20 ···················· ···················· ···················· ···················· ···················· ····················
Q 7754 hCoV-19/England/CAMC-AAF704/2020|EPI_ISL_631224|20 ···················· ···················· ···················· ···················· ···················· ····················
Q 7755 hCoV-19/England/CAMC-AAF5EC/2020|EPI_ISL_631225|20 ···················· ···················· ···················· ···················· ···················· ····················
Q 7756 hCoV-19/Germany/NW-HHU-236/2020|EPI_ISL_631369|202 ···················· ···················· ···················· ···················· ······T············· ····················
Q 7757 hCoV-19/Germany/NW-HHU-237/2020|EPI_ISL_631370|202 ···················· ···················· ···················· ···················· ······T············· ····················
Q 7758 hCoV-19/Germany/NW-HHU-238/2020|EPI_ISL_631371|202 ···················· ···················· ···················· ···················· ······T············· ····················
Q 7759 hCoV-19/Germany/NW-HHU-239/2020|EPI_ISL_631372|202 ···················· ···················· ···················· ···················· ······T············· ····················
Q 7760 hCoV-19/Germany/NW-HHU-240/2020|EPI_ISL_631373|202 ···················· ···················· ···················· ···················· ······T············· ····················
Q 7761 hCoV-19/Germany/NW-HHU-241/2020|EPI_ISL_631374|202 ···················· ···················· ···················· ···················· ······T············· ····················
Q 7762 hCoV-19/Germany/NW-HHU-242/2020|EPI_ISL_631375|202 ···················· ···················· ···················· ···················· ···················· ····················
Q 7763 hCoV-19/Germany/NW-HHU-243/2020|EPI_ISL_631376|202 ···················· ···················· ···················· ···················· ······T············· ····················
Q 7764 hCoV-19/Germany/NW-HHU-244/2020|EPI_ISL_631377|202 ···················· ······A············· ···················· ···················· ···················· ····················
Q 7765 hCoV-19/Germany/NW-HHU-245/2020|EPI_ISL_631378|202 ···················· ···················· ···················· ···················· ···················· ····················
Q 7766 hCoV-19/Germany/NW-HHU-246/2020|EPI_ISL_631379|202 ···················· ···················· ···················· ···················· ···················· ····················
Q 7777 hCoV-19/Netherlands/ZH-EMC-688/2020|EPI_ISL_632542 ···················· ···················· ···················· ···················· ···················· ····················
Q 7778 hCoV-19/Netherlands/ZH-EMC-690/2020|EPI_ISL_632544 ···················· ···················· ···················· ···················· ···················· ····················
Q 7779 hCoV-19/Netherlands/ZH-EMC-691/2020|EPI_ISL_632545 ···················· ···················· ···················· ···················· ···················· ····················
Q 7780 hCoV-19/Netherlands/ZH-EMC-692/2020|EPI_ISL_632546 ···················· ···················· ···················· ···················· ···················· ····················
Q 7781 hCoV-19/Netherlands/ZH-EMC-695/2020|EPI_ISL_632547 ···················· ···················· ···················· ···················· ···················· ····················
Q 7782 hCoV-19/Netherlands/ZH-EMC-696/2020|EPI_ISL_632548 ···················· ···················· ···················· ···················· ···················· ····················
Q 7783 hCoV-19/Netherlands/ZH-EMC-699/2020|EPI_ISL_632549 ···················· ···················· ···················· ···················· ···················· ····················
Q 7784 hCoV-19/Netherlands/ZH-EMC-702/2020|EPI_ISL_632550 ···················· ···················· ···················· ···················· ···················· ····················
Q 7785 hCoV-19/Netherlands/ZH-EMC-703/2020|EPI_ISL_632551 ···················· ···················· ···················· ···················· ···················· ····················
Q 7786 hCoV-19/Netherlands/ZH-EMC-704/2020|EPI_ISL_632552 ···················· ···················· ···················· ···················· ···················· ····················
Q 7787 hCoV-19/Netherlands/ZH-EMC-705/2020|EPI_ISL_632553 ···················· ···················· ···················· ···················· ······T············· ····················
Q 7788 hCoV-19/Netherlands/ZH-EMC-706/2020|EPI_ISL_632554 ···················· ···················· ···················· ···················· ···················· ····················
Q 7789 hCoV-19/Netherlands/ZH-EMC-707/2020|EPI_ISL_632555 ···················· ···················· ···················· ···················· ···················· ····················
Q 7790 hCoV-19/Netherlands/ZH-EMC-708/2020|EPI_ISL_632556 ···················· ···················· ···················· ···················· ···················· ····················
Q 7791 hCoV-19/Netherlands/ZH-EMC-763/2020|EPI_ISL_632600 ···················· ···················· ···················· ···················· ···················· ····················
Q 7792 hCoV-19/Netherlands/ZH-EMC-764/2020|EPI_ISL_632601 ···················· ···················· ···················· ···················· ···················· ····················
Q 7793 hCoV-19/Netherlands/ZH-EMC-765/2020|EPI_ISL_632602 ···················· ···················· ···················· ···················· ···················· ····················
Q 7794 hCoV-19/Netherlands/ZH-EMC-767/2020|EPI_ISL_632603 ···················· ···················· ···················· ···················· ···················· ····················
Q 7795 hCoV-19/Netherlands/ZH-EMC-768/2020|EPI_ISL_632604 ···················· ···················· ···················· ···················· ···················· ····················
Q 7796 hCoV-19/Netherlands/ZH-EMC-769/2020|EPI_ISL_632605 ···················· ···················· ···················· ···················· ···················· ····················
Q 7797 hCoV-19/Netherlands/ZH-EMC-770/2020|EPI_ISL_632606 ···················· ···················· ···················· ···················· ···················· ····················
Q 7798 hCoV-19/Netherlands/ZH-EMC-771/2020|EPI_ISL_632607 ···················· ···················· ···················· ···················· ···················· ····················
Q 7799 hCoV-19/Netherlands/NB-EMC-327/2020|EPI_ISL_632608 ···················· ···················· ···················· ···················· ···················· ····················
Q 7800 hCoV-19/Netherlands/NB-EMC-328/2020|EPI_ISL_632609 ···················· ···················· ···················· ···················· ···················· ····················
Q 7801 hCoV-19/Netherlands/ZH-EMC-773/2020|EPI_ISL_632610 ···················· ···················· ···················· ···················· ···················· ····················
Q 7802 hCoV-19/Netherlands/ZH-EMC-774/2020|EPI_ISL_632611 ···················· ···················· ···················· ···················· ···················· ····················
Q 7803 hCoV-19/England/QEUH-AD30C0/2020|EPI_ISL_633873|20 ···················· ···················· ···················· ···················· ···················· ····················
Q 7804 hCoV-19/England/QEUH-AD3136/2020|EPI_ISL_633875|20 ···················· ···················· ···················· ···················· ···················· ····················
Q 7805 hCoV-19/England/QEUH-AD27BD/2020|EPI_ISL_633876|20 ···················· ···················· ···················· ···················· ···················· ····················
Q 7806 hCoV-19/England/QEUH-AD5592/2020|EPI_ISL_633879|20 ···················· ···················· ···················· ···················· ···················· ····················
Q 7807 hCoV-19/England/QEUH-AD4FA4/2020|EPI_ISL_633880|20 ···················· ···················· ···················· ···················· ···················· ····················
Q 7808 hCoV-19/England/QEUH-AD0A5C/2020|EPI_ISL_633881|20 ···················· ···················· ···················· ···················· ···················· ····················
Q 7809 hCoV-19/England/QEUH-AD0A10/2020|EPI_ISL_633882|20 ···················· ·A·················· ···················· ···················· ···················· ···A················
Q 7810 hCoV-19/England/QEUH-AD27CC/2020|EPI_ISL_633883|20 ···················· ···················· ···················· ···················· ···················· ····················
Q 7811 hCoV-19/England/QEUH-AD154B/2020|EPI_ISL_633884|20 ···················· ···················· ···················· ···················· ···················· ····················
Q 7812 hCoV-19/England/QEUH-AD0B59/2020|EPI_ISL_633885|20 ···················· ···················· ···················· ···················· ···················· ····················

| | | | | | | | |
|---|---|---|---|---|---|---|---|
| Q | 7813 hCoV-19/England/QEUH-AD0E32/2020\|EPI_ISL_633901\|20 | ···················· | ···················· | ···················· | ···················· | ···················· | ···················· |
| Q | 7814 hCoV-19/England/QEUH-AD387D/2020\|EPI_ISL_633902\|20 | ···················· | ···················· | ···················· | ···················· | ···················· | ···················· |
| Q | 7815 hCoV-19/England/QEUH-AD27DB/2020\|EPI_ISL_633904\|20 | ···················· | ···················· | ···················· | ···················· | ···················· | ···················· |
| Q | 7816 hCoV-19/England/QEUH-AD0B68/2020\|EPI_ISL_633906\|20 | ···················· | ···················· | ···················· | ···················· | ···················· | ···················· |
| Q | 7817 hCoV-19/England/QEUH-AD12BD/2020\|EPI_ISL_633908\|20 | ···················· | ···················· | ···················· | ···················· | ···················· | ···················· |
| Q | 7818 hCoV-19/England/QEUH-AD54FF/2020\|EPI_ISL_633909\|20 | ···················· | ···················· | ···················· | ···················· | ···················· | ···················· |
| Q | 7819 hCoV-19/England/QEUH-AD5529/2020\|EPI_ISL_633911\|20 | ···················· | ···················· | ···················· | ···················· | ···················· | ···················· |
| Q | 7820 hCoV-19/England/QEUH-AD0B4A/2020\|EPI_ISL_633912\|20 | ···················· | ···················· | ···················· | ···················· | ···················· | ···················· |
| Q | 7821 hCoV-19/England/QEUH-AD389B/2020\|EPI_ISL_633913\|20 | ···················· | ···················· | ···················· | ···················· | ···················· | ···················· |
| Q | 7822 hCoV-19/England/QEUH-AD35C1/2020\|EPI_ISL_633915\|20 | ···················· | ···················· | ···················· | ···················· | ···················· | ···················· |
| Q | 7823 hCoV-19/England/QEUH-AD5D30/2020\|EPI_ISL_633917\|20 | ···················· | ···················· | ···················· | ···················· | ···················· | ···················· |
| Q | 7824 hCoV-19/England/QEUH-AD5028/2020\|EPI_ISL_633918\|20 | ···················· | ···················· | ···················· | ···················· | ···················· | ···················· |
| Q | 7825 hCoV-19/England/QEUH-AD2832/2020\|EPI_ISL_633919\|20 | ···················· | ···················· | ···················· | ···················· | ···················· | ···················· |
| Q | 7826 hCoV-19/England/QEUH-AD5DE5/2020\|EPI_ISL_633920\|20 | ···················· | ···················· | ···················· | ···················· | ···················· | ···················· |
| Q | 7827 hCoV-19/England/QEUH-AD153C/2020\|EPI_ISL_633921\|20 | ···················· | ···················· | ···················· | ···················· | ···················· | ···················· |
| Q | 7828 hCoV-19/England/QEUH-AD1B2B/2020\|EPI_ISL_633924\|20 | ···················· | ···················· | ···················· | ···················· | ···················· | ···················· |
| Q | 7829 hCoV-19/England/QEUH-AD3673/2020\|EPI_ISL_633925\|20 | ···················· | ···················· | ···················· | ···················· | ···················· | ···················· |
| Q | 7830 hCoV-19/England/QEUH-AD5538/2020\|EPI_ISL_633926\|20 | ···················· | ·A·················· | ···················· | ···················· | ···················· | ···················· |
| Q | 7831 hCoV-19/England/QEUH-AD27F9/2020\|EPI_ISL_633927\|20 | ···················· | ·A·················· | ···················· | ···················· | ···················· | ···················· |
| Q | 7832 hCoV-19/England/QEUH-AD0B77/2020\|EPI_ISL_633928\|20 | ···················· | ···················· | ···················· | ···················· | ···················· | ···················· |
| Q | 7833 hCoV-19/England/QEUH-AD288D/2020\|EPI_ISL_633929\|20 | ···················· | ···················· | ···················· | ···················· | ···················· | ···················· |
| Q | 7834 hCoV-19/England/QEUH-AD590F/2020\|EPI_ISL_633930\|20 | ···················· | ···················· | ···················· | ···················· | ···················· | ···················· |
| Q | 7835 hCoV-19/England/QEUH-AD3585/2020\|EPI_ISL_633931\|20 | ···················· | ···················· | ···················· | ···················· | ···················· | ···················· |
| Q | 7836 hCoV-19/England/QEUH-AD36A0/2020\|EPI_ISL_633932\|20 | ···················· | ···················· | ···················· | ···················· | ···················· | ···················· |
| Q | 7837 hCoV-19/England/QEUH-AD59A5/2020\|EPI_ISL_633933\|20 | ···················· | ···················· | ···················· | ···················· | ···················· | ···················· |
| Q | 7838 hCoV-19/Wales/QEUH-AD32E8/2020\|EPI_ISL_633934\|2020 | ···················· | ···················· | ···················· | ···················· | ···················· | ···················· |
| Q | 7839 hCoV-19/England/QEUH-AD4E6B/2020\|EPI_ISL_633935\|20 | ···················· | ···················· | ···················· | ···················· | ···················· | ···················· |
| Q | 7840 hCoV-19/England/QEUH-AD334F/2020\|EPI_ISL_633936\|20 | ···················· | ···················· | ···················· | ···················· | ···················· | ···················· |
| Q | 7841 hCoV-19/England/QEUH-AD0E7E/2020\|EPI_ISL_633938\|20 | ···················· | ···················· | ···················· | ···················· | ···················· | ···················· |
| Q | 7842 hCoV-19/England/QEUH-AD155A/2020\|EPI_ISL_633939\|20 | ···················· | ···················· | ···················· | ···················· | ······T············· | ···················· |
| Q | 7843 hCoV-19/England/QEUH-AD1174/2020\|EPI_ISL_633940\|20 | ···················· | ···················· | ···················· | ···················· | ···················· | ···················· |
| Q | 7844 hCoV-19/England/QEUH-AD397A/2020\|EPI_ISL_633941\|20 | ···················· | ···················· | ···················· | ···················· | ···················· | ···················· |
| Q | 7845 hCoV-19/England/QEUH-AD329D/2020\|EPI_ISL_633942\|20 | ···················· | ···················· | ···················· | ···················· | ···················· | ···················· |
| Q | 7846 hCoV-19/USA/CA-ALSR-3593/2020\|EPI_ISL_636100\|2020- | ···················· | ·A·················· | ···················· | ···················· | ···················· | ···················· |
| Q | 7847 hCoV-19/USA/CA-ALSR-3595/2020\|EPI_ISL_636101\|2020- | ···················· | ·A·················· | ···················· | ···················· | ···················· | ···················· |
| Q | 7848 hCoV-19/USA/CA-ALSR-3596/2020\|EPI_ISL_636102\|2020- | ···················· | ·A·················· | ···················· | ···················· | ···················· | ···················· |
| Q | 7849 hCoV-19/USA/CA-ALSR-3598/2020\|EPI_ISL_636104\|2020- | ···················· | ···················· | ···················· | ···················· | ···················· | ···················· |
| Q | 7850 hCoV-19/USA/CA-ALSR-3600/2020\|EPI_ISL_636105\|2020- | ···················· | ·A·················· | ···················· | ···················· | ···················· | ···················· |
| Q | 7851 hCoV-19/USA/CA-ALSR-3601/2020\|EPI_ISL_636106\|2020- | ···················· | ·A·················· | ···················· | ···················· | ···················· | ···················· |
| Q | 7852 hCoV-19/USA/CA-ALSR-3604/2020\|EPI_ISL_636107\|2020- | ···················· | ···················· | ···················· | ···················· | ···················· | ···················· |
| Q | 7853 hCoV-19/USA/CA-ALSR-3606/2020\|EPI_ISL_636108\|2020- | ···················· | ·A·················· | ···················· | ···················· | ···················· | ···················· |
| Q | 7854 hCoV-19/USA/CA-ALSR-3609/2020\|EPI_ISL_636109\|2020- | ···················· | ·A·················· | ···················· | ···················· | ···················· | ···················· |
| Q | 7855 hCoV-19/USA/CA-ALSR-3610/2020\|EPI_ISL_636110\|2020- | ···················· | ·A·················· | ···················· | ···················· | ···················· | ···················· |
| Q | 7856 hCoV-19/USA/CA-ALSR-3612/2020\|EPI_ISL_636111\|2020- | ···················· | ·A·················· | ···················· | ···················· | ···················· | ···················· |
| Q | 7857 hCoV-19/USA/CA-ALSR-3614/2020\|EPI_ISL_636112\|2020- | ···················· | ·A·················· | ···················· | ···················· | ···················· | ···················· |
| Q | 7858 hCoV-19/Mexico/BCN-ALSR-3618/2020\|EPI_ISL_636113\|2 | ···················· | ·A·················· | ···················· | ···················· | ···················· | ···················· |
| Q | 7859 hCoV-19/Mexico/BCN-ALSR-3619/2020\|EPI_ISL_636114\|2 | ···················· | ·A·················· | ···················· | ···················· | ···················· | ···················· |
| Q | 7860 hCoV-19/Mexico/BCN-ALSR-3622/2020\|EPI_ISL_636115\|2 | ···················· | ·A·················· | ···················· | ···················· | ···················· | ···················· |
| Q | 7861 hCoV-19/Mexico/BCN-ALSR-3624/2020\|EPI_ISL_636116\|2 | ···················· | ·A·················· | ···················· | ···················· | ···················· | ···················· |
| Q | 7862 hCoV-19/Mexico/BCN-ALSR-3630/2020\|EPI_ISL_636117\|2 | ···················· | ·A·················· | ···················· | ···················· | ···················· | ···················· |
| Q | 7863 hCoV-19/Mexico/BCN-ALSR-3634/2020\|EPI_ISL_636118\|2 | ···················· | ·A·················· | ···················· | ···················· | ···················· | ···················· |
| Q | 7864 hCoV-19/Mexico/BCN-ALSR-3636/2020\|EPI_ISL_636119\|2 | ···················· | ·A·················· | ···················· | ···················· | ···················· | ···················· |
| Q | 7865 hCoV-19/USA/CA-ALSR-3643/2020\|EPI_ISL_636121\|2020- | ········T··········· | ·A·················· | ···················· | ···················· | ···················· | ···················· |
| Q | 7866 hCoV-19/USA/CA-ALSR-3644/2020\|EPI_ISL_636122\|2020- | ···················· | ·A·················· | ···················· | ···················· | ···················· | ···················· |
| Q | 7867 hCoV-19/USA/CA-ALSR-3645/2020\|EPI_ISL_636123\|2020- | ···················· | ·A·················· | ···················· | ···················· | ···················· | ···················· |
| Q | 7868 hCoV-19/USA/CA-ALSR-3646/2020\|EPI_ISL_636124\|2020- | ···················· | ···················· | ···················· | ···················· | ···················· | ···················· |
| Q | 7869 hCoV-19/USA/CA-ALSR-3647/2020\|EPI_ISL_636125\|2020- | ···················· | ···················· | ···················· | ···················· | ···················· | ···················· |
| Q | 7870 hCoV-19/USA/CA-ALSR-3648/2020\|EPI_ISL_636126\|2020- | ···················· | ·A·················· | ···················· | ···················· | ···················· | ···················· |
| Q | 7871 hCoV-19/USA/CA-ALSR-3650/2020\|EPI_ISL_636127\|2020- | ···················· | ·A·················· | ···················· | ···················· | ···················· | ···················· |
| Q | 7872 hCoV-19/USA/CA-ALSR-3651/2020\|EPI_ISL_636128\|2020- | ···················· | ·A·················· | ···················· | ···················· | ···················· | ···················· |
| Q | 7873 hCoV-19/USA/CA-ALSR-3653/2020\|EPI_ISL_636129\|2020- | ···················· | ···················· | ···················· | ···················· | ···················· | ···················· |
| Q | 7874 hCoV-19/USA/CA-ALSR-3655/2020\|EPI_ISL_636130\|2020- | ···················· | ·A·················· | ···················· | ···················· | ···················· | ···················· |
| Q | 7875 hCoV-19/USA/CA-ALSR-3658/2020\|EPI_ISL_636131\|2020- | ···················· | ···················· | ···················· | ···················· | ···················· | ···················· |
| Q | 7876 hCoV-19/USA/CA-ALSR-3659/2020\|EPI_ISL_636132\|2020- | ···················· | ·A·················· | ···················· | ···················· | ···················· | ···················· |
| Q | 7877 hCoV-19/USA/CA-ALSR-3660/2020\|EPI_ISL_636133\|2020- | ···················· | ·A·················· | ···················· | ···················· | ···················· | ···················· |
| Q | 7878 hCoV-19/USA/CA-ALSR-3661/2020\|EPI_ISL_636134\|2020- | ···················· | ···················· | ···················· | ···················· | ···················· | ···················· |
| Q | 7879 hCoV-19/USA/CA-ALSR-3662/2020\|EPI_ISL_636135\|2020- | ···················· | ·A·················· | ···················· | ···················· | ···················· | ···················· |
| Q | 7880 hCoV-19/USA/CA-ALSR-3664/2020\|EPI_ISL_636136\|2020- | ···················· | ···················· | ···················· | ···················· | ···················· | ···················· |
| Q | 7881 hCoV-19/USA/CA-ALSR-3666/2020\|EPI_ISL_636137\|2020- | ···················· | ·A·················· | ···················· | ···················· | ···················· | ···················· |
| Q | 7882 hCoV-19/USA/CA-ALSR-3667/2020\|EPI_ISL_636138\|2020- | ···················· | ···················· | ···················· | ···················· | ···················· | ···················· |
| Q | 7883 hCoV-19/USA/CA-ALSR-3668/2020\|EPI_ISL_636139\|2020- | ···················· | ·A·················· | ···················· | ···················· | ···················· | ···················· |
| Q | 7884 hCoV-19/USA/CA-ALSR-3669/2020\|EPI_ISL_636140\|2020- | ···················· | ·A·················· | ···················· | ···················· | ···················· | ···················· |
| Q | 7885 hCoV-19/USA/CA-ALSR-3670/2020\|EPI_ISL_636141\|2020- | ···················· | ···················· | ···················· | ···················· | ···················· | ···················· |
| Q | 7886 hCoV-19/USA/CA-ALSR-3676/2020\|EPI_ISL_636143\|2020- | ···················· | ·A·················· | ···················· | ···················· | ···················· | ···················· |
| Q | 7887 hCoV-19/USA/CA-ALSR-3678/2020\|EPI_ISL_636144\|2020- | ···················· | ·A·················· | ···················· | ···················· | ···················· | ···················· |
| Q | 7888 hCoV-19/USA/CA-ALSR-3680/2020\|EPI_ISL_636145\|2020- | ···················· | ···················· | ···················· | ···················· | ···················· | ···················· |
| Q | 7889 hCoV-19/USA/CA-ALSR-3684/2020\|EPI_ISL_636146\|2020- | ···················· | ·A·················· | ···················· | ···················· | ···················· | ···················· |
| Q | 7890 hCoV-19/USA/CA-ALSR-3685/2020\|EPI_ISL_636147\|2020- | ···················· | ·A·················· | ···················· | ···················· | ···················· | ···················· |
| Q | 7891 hCoV-19/USA/CA-ALSR-3686/2020\|EPI_ISL_636148\|2020- | ···················· | ·A·················· | ···················· | ···················· | ···················· | ···················· |
| Q | 7892 hCoV-19/USA/CA-ALSR-3688/2020\|EPI_ISL_636149\|2020- | ···················· | ·A·················· | ···················· | ···················· | ···················· | ···················· |

| | | | | | | | |
|---|---|---|---|---|---|---|---|
| Q | 7893 hCoV-19/USA/CA-ALSR-3691/2020\|EPI_ISL_636150\|2020- | ···················· | ·A·················· | ····················· | ···················· | ···················· | ···················· |
| Q | 7894 hCoV-19/USA/CA-ALSR-3693/2020\|EPI_ISL_636151\|2020- | ···················· | ·A·················· | ····················· | ···················· | ···················· | ···················· |
| Q | 7895 hCoV-19/USA/CA-ALSR-3694/2020\|EPI_ISL_636152\|2020- | ···················· | ···················· | ····················· | ···················· | ···················· | ···················· |
| Q | 7896 hCoV-19/USA/CA-ALSR-3699/2020\|EPI_ISL_636153\|2020- | ···················· | ·A·················· | ····················· | ···················· | ···················· | ···················· |
| Q | 7897 hCoV-19/USA/CA-ALSR-3700/2020\|EPI_ISL_636154\|2020- | ···················· | ···················· | ····················· | ···················· | ···················· | ···················· |
| Q | 7898 hCoV-19/USA/CA-ALSR-3705/2020\|EPI_ISL_636156\|2020- | ···················· | ···················· | ····················· | ···················· | ···················· | ···················· |
| Q | 7899 hCoV-19/USA/CA-ALSR-3706/2020\|EPI_ISL_636157\|2020- | ···················· | ·A·················· | ····················· | ···················· | ···················· | ···················· |
| Q | 7900 hCoV-19/USA/CA-ALSR-3707/2020\|EPI_ISL_636158\|2020- | ·······T············ | ·A·················· | ····················· | ···················· | ···················· | ···················· |
| Q | 7901 hCoV-19/USA/CA-ALSR-3713/2020\|EPI_ISL_636160\|2020- | ···················· | ·A·················· | ····················· | ···················· | ···················· | ···················· |
| Q | 7902 hCoV-19/USA/CA-ALSR-3714/2020\|EPI_ISL_636161\|2020- | ···················· | ···················· | ····················· | ···················· | ···················· | ···················· |
| Q | 7903 hCoV-19/USA/CA-ALSR-3716/2020\|EPI_ISL_636162\|2020- | ···················· | ···················· | ····················· | ···················· | ···················· | ···················· |
| Q | 7904 hCoV-19/USA/CA-ALSR-3717/2020\|EPI_ISL_636163\|2020- | ···················· | ·A·················· | ····················· | ···················· | ···················· | ···················· |
| Q | 7905 hCoV-19/USA/CA-ALSR-3721/2020\|EPI_ISL_636165\|2020- | ···················· | ·A·················· | ····················· | ···················· | ···················· | ···················· |
| Q | 7906 hCoV-19/USA/CA-ALSR-3722/2020\|EPI_ISL_636166\|2020- | ···················· | ·A·················· | ····················· | ···················· | ···················· | ···················· |
| Q | 7907 hCoV-19/USA/CA-ALSR-3723/2020\|EPI_ISL_636167\|2020- | ···················· | ·A·················· | ····················· | ···················· | ···················· | ···················· |
| Q | 7908 hCoV-19/USA/CA-ALSR-3725/2020\|EPI_ISL_636168\|2020- | ···················· | ·A·················· | ····················· | ···················· | ···················· | ···················· |
| Q | 7909 hCoV-19/USA/CA-ALSR-3726/2020\|EPI_ISL_636169\|2020- | ···················· | ···················· | ····················· | ···················· | ···················· | ···················· |
| Q | 7910 hCoV-19/USA/CA-ALSR-3728/2020\|EPI_ISL_636170\|2020- | ···················· | ···················· | ····················· | ···················· | ···················· | ···················· |
| Q | 7911 hCoV-19/USA/CA-ALSR-3731/2020\|EPI_ISL_636172\|2020- | ···················· | ·A·················· | ····················· | ···················· | ···················· | ···················· |
| Q | 7912 hCoV-19/USA/CA-ALSR-3732/2020\|EPI_ISL_636173\|2020- | ···················· | ·A·················· | ····················· | ···················· | ···················· | ···················· |
| Q | 7913 hCoV-19/USA/CA-ALSR-3736/2020\|EPI_ISL_636174\|2020- | ···················· | ·A·················· | ····················· | ···················· | ···················· | ···················· |
| Q | 7914 hCoV-19/USA/CA-ALSR-3737/2020\|EPI_ISL_636175\|2020- | ···················· | ·A·················· | ····················· | ···················· | ···················· | ···················· |
| Q | 7915 hCoV-19/USA/CA-ALSR-3739/2020\|EPI_ISL_636177\|2020- | ···················· | ·A·················· | ····················· | ···················· | ···················· | ···················· |
| Q | 7916 hCoV-19/USA/CA-ALSR-3741/2020\|EPI_ISL_636178\|2020- | ···················· | ·A·················· | ····················· | ···················· | ···················· | ···················· |
| Q | 7917 hCoV-19/USA/CA-ALSR-3747/2020\|EPI_ISL_636179\|2020- | ···················· | ·A·················· | ····················· | ···················· | ···················· | ···················· |
| Q | 7918 hCoV-19/USA/CA-ALSR-3749/2020\|EPI_ISL_636180\|2020- | ···················· | ·A·················· | ····················· | ···················· | ···················· | ···················· |
| Q | 7919 hCoV-19/USA/CA-ALSR-3751/2020\|EPI_ISL_636181\|2020- | ···················· | ·A·················· | ····················· | ···················· | ···················· | ···················· |
| Q | 7920 hCoV-19/USA/CA-ALSR-3752/2020\|EPI_ISL_636182\|2020- | ·······T············ | ···················· | ····················· | ···················· | ···················· | ···················· |
| Q | 7921 hCoV-19/USA/CA-ALSR-3753/2020\|EPI_ISL_636183\|2020- | ···················· | ···················· | ····················· | ···················· | ···················· | ···················· |
| Q | 7922 hCoV-19/USA/CA-ALSR-3754/2020\|EPI_ISL_636184\|2020- | ···················· | ·A·················· | ····················· | ···················· | ···················· | ···················· |
| Q | 7923 hCoV-19/USA/CA-ALSR-3756/2020\|EPI_ISL_636185\|2020- | ···················· | ·A·················· | ····················· | ···················· | ···················· | ···················· |
| Q | 7924 hCoV-19/USA/CA-ALSR-3759/2020\|EPI_ISL_636186\|2020- | ···················· | ·A·················· | ····················· | ···················· | ···················· | ···················· |
| Q | 7925 hCoV-19/USA/CA-ALSR-3760/2020\|EPI_ISL_636187\|2020- | ···················· | ·A·················· | ····················· | ···················· | ···················· | ···················· |
| Q | 7926 hCoV-19/USA/CA-ALSR-3761/2020\|EPI_ISL_636188\|2020- | ···················· | ·A·················· | ····················· | ···················· | ···················· | ···················· |
| Q | 7927 hCoV-19/USA/CA-ALSR-3764/2020\|EPI_ISL_636189\|2020- | ···················· | ·A·················· | ····················· | ···················· | ···················· | ···················· |
| Q | 7928 hCoV-19/USA/CA-ALSR-3770/2020\|EPI_ISL_636190\|2020- | ···················· | ·A·················· | ····················· | ···················· | ···················· | ···················· |
| Q | 7929 hCoV-19/USA/CA-ALSR-3773/2020\|EPI_ISL_636192\|2020- | ···················· | ·A·················· | ····················· | ···················· | ···················· | ···················· |
| Q | 7930 hCoV-19/USA/CA-ALSR-3774/2020\|EPI_ISL_636193\|2020- | ···················· | ·A·················· | ····················· | ···················· | ···················· | ···················· |
| Q | 7931 hCoV-19/USA/CA-ALSR-3776/2020\|EPI_ISL_636194\|2020- | ···················· | ·A·················· | ····················· | ···················· | ···················· | ···················· |
| Q | 7932 hCoV-19/USA/CA-ALSR-3777/2020\|EPI_ISL_636195\|2020- | ···················· | ·A·················· | ····················· | ···················· | ···················· | ···················· |
| Q | 7933 hCoV-19/USA/CA-ALSR-3780/2020\|EPI_ISL_636196\|2020- | ···················· | ·A·················· | ····················· | ···················· | ···················· | ···················· |
| Q | 7934 hCoV-19/USA/CA-ALSR-3781/2020\|EPI_ISL_636197\|2020- | ···················· | ·A·················· | ····················· | ···················· | ···················· | ···················· |
| Q | 7935 hCoV-19/USA/CA-ALSR-3782/2020\|EPI_ISL_636198\|2020- | ···················· | ·A·················· | ····················· | ···················· | ···················· | ···················· |
| Q | 7936 hCoV-19/USA/CA-ALSR-3784/2020\|EPI_ISL_636199\|2020- | ···················· | ·A·················· | ····················· | ···················· | ···················· | ···················· |
| Q | 7937 hCoV-19/Singapore/1117/2020\|EPI_ISL_626628\|2020-10 | ···················· | ···················· | ····················· | ···················· | ···················· | ···················· |
| Q | 7938 hCoV-19/Singapore/1118/2020\|EPI_ISL_626629\|2020-10 | ···················· | ·A·················· | ····················· | ···················· | ···················· | ···················· |
| Q | 7939 hCoV-19/Singapore/1120/2020\|EPI_ISL_626631\|2020-10 | ···················· | ·········A·········· | ····················· | ···················· | ···················· | ···················· |
| Q | 7940 hCoV-19/Singapore/1121/2020\|EPI_ISL_626632\|2020-10 | ···················· | ···················· | ····················· | ···················· | ···················· | ···················· |
| Q | 7941 hCoV-19/Singapore/1122/2020\|EPI_ISL_626633\|2020-10 | ···················· | ···················· | ····················· | ···················· | ···················· | ···················· |
| Q | 7942 hCoV-19/Singapore/1123/2020\|EPI_ISL_626634\|2020-10 | ···················· | ···················· | ····················· | ···················· | ···················· | ···················· |
| Q | 7943 hCoV-19/Wales/PHWC-48405B/2020\|EPI_ISL_627959\|2020 | ···················· | ···················· | ····················· | ···················· | ···················· | ···················· |
| Q | 7944 hCoV-19/Wales/PHWC-484097/2020\|EPI_ISL_627962\|2020 | ···················· | ···················· | ····················· | ···················· | ···················· | ···················· |
| Q | 7945 hCoV-19/Wales/PHWC-4840B5/2020\|EPI_ISL_627963\|2020 | ···················· | ···················· | ····················· | ···················· | ···················· | ···················· |
| Q | 7946 hCoV-19/Wales/PHWC-4840D3/2020\|EPI_ISL_627964\|2020 | ···················· | ···················· | ····················· | ···················· | ···················· | ···················· |
| Q | 7948 hCoV-19/Wales/PHWC-48412B/2020\|EPI_ISL_627968\|2020 | ···················· | ···················· | ····················· | ···················· | ···················· | ···················· |
| Q | 7949 hCoV-19/Wales/PHWC-484325/2020\|EPI_ISL_627990\|2020 | ···················· | ···················· | ····················· | ···················· | ···················· | ···················· |
| Q | 7950 hCoV-19/Wales/PHWC-484334/2020\|EPI_ISL_627991\|2020 | ···················· | ···················· | ····················· | ···················· | ···················· | ···················· |
| Q | 7951 hCoV-19/Wales/PHWC-484343/2020\|EPI_ISL_627992\|2020 | ···················· | ···················· | ····················· | ···················· | ···················· | ···················· |
| Q | 7952 hCoV-19/Wales/PHWC-484352/2020\|EPI_ISL_627993\|2020 | ···················· | ···················· | ····················· | ···················· | ···················· | ···················· |
| Q | 7953 hCoV-19/Wales/PHWC-484370/2020\|EPI_ISL_627994\|2020 | ···················· | ···················· | ····················· | ···················· | ···················· | ···················· |
| Q | 7954 hCoV-19/Wales/PHWC-48438F/2020\|EPI_ISL_627995\|2020 | ···················· | ···················· | ····················· | ···················· | ···················· | ···················· |
| Q | 7955 hCoV-19/Wales/PHWC-48439E/2020\|EPI_ISL_627996\|2020 | ···················· | ···················· | ····················· | ···················· | ···················· | ···················· |
| Q | 7956 hCoV-19/Wales/PHWC-4843AD/2020\|EPI_ISL_627997\|2020 | ···················· | ···················· | ····················· | ···················· | ···················· | ···················· |
| Q | 7957 hCoV-19/Wales/PHWC-4843BC/2020\|EPI_ISL_627998\|2020 | ···················· | ···················· | ····················· | ···················· | ···················· | ···················· |
| Q | 7958 hCoV-19/Wales/PHWC-4843CB/2020\|EPI_ISL_627999\|2020 | ···················· | ···················· | ····················· | ···················· | ···················· | ···················· |
| Q | 7959 hCoV-19/England/MILK-ABBEB5/2020\|EPI_ISL_631200\|20 | ···················· | ···················· | ····················· | ···················· | ···················· | ···················· |
| Q | 7960 hCoV-19/England/MILK-ABBC42/2020\|EPI_ISL_631201\|20 | ···················· | ···················· | ····················· | ···················· | ···················· | ···················· |
| Q | 7961 hCoV-19/England/ALDP-ABD220/2020\|EPI_ISL_631202\|20 | ···················· | ···················· | ····················· | ···················· | ···················· | ···················· |
| Q | 7962 hCoV-19/England/CAMC-AAF8F2/2020\|EPI_ISL_631204\|20 | ···················· | ···················· | ····················· | ···················· | ···················· | ···················· |
| Q | 7963 hCoV-19/England/MILK-ABBF58/2020\|EPI_ISL_631205\|20 | ···················· | ···················· | ····················· | ···················· | ···················· | ···················· |
| Q | 7964 hCoV-19/England/MILK-ABC33D/2020\|EPI_ISL_631206\|20 | ···················· | ···················· | ····················· | ···················· | ······T············· | ···················· |
| Q | 7965 hCoV-19/England/MILK-ABC4EF/2020\|EPI_ISL_631207\|20 | ···················· | ···················· | ····················· | ···················· | ···················· | ···················· |
| Q | 7966 hCoV-19/England/CAMC-AAF713/2020\|EPI_ISL_631208\|20 | ···················· | ···················· | ····················· | ···················· | ···················· | ···················· |
| Q | 7967 hCoV-19/England/MILK-ABC19D/2020\|EPI_ISL_631209\|20 | ···················· | ···················· | ····················· | ···················· | ···················· | ···················· |
| Q | 7968 hCoV-19/England/MILK-ABC160/2020\|EPI_ISL_631210\|20 | ···················· | ···················· | ····················· | ···················· | ···················· | ···················· |
| Q | 7969 hCoV-19/England/MILK-ABBD9A/2020\|EPI_ISL_631213\|20 | ···················· | ···················· | ····················· | ···················· | ···················· | ···················· |
| Q | 7970 hCoV-19/England/MILK-ABBB18/2020\|EPI_ISL_631226\|20 | ···················· | ···················· | ····················· | ···················· | ···················· | ···················· |
| Q | 7971 hCoV-19/England/MILK-ABB565/2020\|EPI_ISL_631227\|20 | ···················· | ···················· | ····················· | ···················· | ···················· | ···················· |
| Q | 7972 hCoV-19/England/ALDP-ABD299/2020\|EPI_ISL_631228\|20 | ···················· | ···················· | ····················· | ···················· | ···················· | ···················· |
| Q | 7973 hCoV-19/England/MILK-ABBB36/2020\|EPI_ISL_631229\|20 | ···················· | ···················· | ····················· | ···················· | ···················· | ···················· |

```
Q  7974 hCoV-19/England/MILK-ABBF94/2020|EPI_ISL_631230|20   ···················· ···················· ···················· ···················· ···················· ····················
Q  7975 hCoV-19/England/ALDP-AC121A/2020|EPI_ISL_631231|20   ···················· ···················· ···················· ···················· ···················· ····················
Q  7976 hCoV-19/England/MILK-ABC485/2020|EPI_ISL_631232|20   ···················· ·A·················· ···················· ···················· ···················· ··A·················
Q  7977 hCoV-19/England/MILK-ABC2C7/2020|EPI_ISL_631233|20   ···················· ···················· ···················· ···················· ·····T·············· ····················
Q  7978 hCoV-19/England/MILK-ABBBAF/2020|EPI_ISL_631234|20   ···················· ···················· ···················· ···················· ···················· ····················
Q  7979 hCoV-19/England/CAMC-AAF77D/2020|EPI_ISL_631235|20   ···················· ···················· ···················· ···················· ···················· ····················
Q  7980 hCoV-19/England/MILK-ABC0AF/2020|EPI_ISL_631236|20   ···················· ···················· ···················· ···················· ···················· ····················
Q  7981 hCoV-19/England/CAMC-AAF9A4/2020|EPI_ISL_631237|20   ···················· ···················· ···················· ···················· ···················· ····················
Q  7982 hCoV-19/England/MILK-ABC0FA/2020|EPI_ISL_631238|20   ···················· ···················· ···················· ···················· ···················· ····················
Q  7983 hCoV-19/England/MILK-ABBDA9/2020|EPI_ISL_631240|20   ···················· ···················· ···················· ···················· ···················· ····················
Q  7984 hCoV-19/England/ALDP-ABD28A/2020|EPI_ISL_631241|20   ···················· ···················· ···················· ···················· ···················· ····················
Q  7985 hCoV-19/England/MILK-ABBFA3/2020|EPI_ISL_631242|20   ···················· ···················· ···················· ···················· ···················· ····················
Q  7986 hCoV-19/England/MILK-ABC35B/2020|EPI_ISL_631243|20   ···················· ···················· ···················· ···················· ···················· ····················
Q  7987 hCoV-19/England/MILK-ABBEA6/2020|EPI_ISL_631244|20   ···················· ···················· ···················· ···················· ···················· ····················
Q  7988 hCoV-19/England/CAMC-AAF92C/2020|EPI_ISL_631245|20   ···················· ···················· ···················· ···················· ···················· ····················
Q  7989 hCoV-19/England/MILK-ABBC8E/2020|EPI_ISL_631246|20   ···················· ···················· ···················· ···················· ···················· ····················
Q  7990 hCoV-19/England/MILK-ABBCF7/2020|EPI_ISL_631247|20   ···················· ···················· ···················· ···················· ···················· ····················
Q  7991 hCoV-19/England/MILK-ABBE6A/2020|EPI_ISL_631248|20   ···················· ···················· ···················· ···················· ·····T·············· ····················
Q  7992 hCoV-19/England/MILK-ABB5B0/2020|EPI_ISL_631249|20   ···················· ···················· ···················· ···················· ···················· ····················
Q  7993 hCoV-19/England/ALDP-AC5E21/2020|EPI_ISL_631250|20   ···················· ···················· ···················· ···················· ···················· ····················
Q  7994 hCoV-19/England/MILK-ABB88A/2020|EPI_ISL_631251|20   ···················· ···················· ···················· ···················· ···················· ····················
Q  7995 hCoV-19/England/MILK-ABBCAC/2020|EPI_ISL_631252|20   ···················· ···················· ···················· ···················· ···················· ····················
Q  7996 hCoV-19/England/MILK-ABC3B5/2020|EPI_ISL_631253|20   ···················· ···················· ···················· ···················· ···················· ····················
Q  7997 hCoV-19/England/CAMC-AAF7C8/2020|EPI_ISL_631254|20   ···················· ···················· ···················· ···················· ···················· ····················
Q  7998 hCoV-19/England/MILK-ABC1BB/2020|EPI_ISL_631255|20   ···················· ···················· ···················· ···················· ·····T·············· ····················
Q  7999 hCoV-19/England/MILK-ABB899/2020|EPI_ISL_631256|20   ···················· ···················· ···················· ···················· ···················· ····················
Q  8000 hCoV-19/England/CAMC-AAFF75/2020|EPI_ISL_631257|20   ···················· ···················· ···················· ···················· ···················· ····················
Q  8001 hCoV-19/England/MILK-ABBAFD/2020|EPI_ISL_631258|20   ···················· ···················· ···················· ···················· ···················· ····················
Q  8002 hCoV-19/England/MILK-ABBC60/2020|EPI_ISL_631259|20   ···················· ·A·················· ···················· ···················· ···················· ··A·················
Q  8003 hCoV-19/England/MILK-ABC106/2020|EPI_ISL_631260|20   ···················· ···················· ···················· ···················· ···················· ····················
Q  8004 hCoV-19/England/MILK-ABBEE2/2020|EPI_ISL_631261|20   ···················· ···················· ···················· ···················· ···················· ····················
Q  8005 hCoV-19/England/CAMC-AAF8B6/2020|EPI_ISL_631262|20   ···················· ···················· ···················· ···················· ···················· ····················
Q  8006 hCoV-19/England/MILK-ABC124/2020|EPI_ISL_631264|20   ···················· ···················· ···················· ···················· ···················· ····················
Q  8007 hCoV-19/England/ALDP-AC5D9D/2020|EPI_ISL_631265|20   ···················· ···················· ···················· ···················· ···················· ····················
Q  8008 hCoV-19/England/CAMC-AAF8C5/2020|EPI_ISL_631266|20   ···················· ···················· ···················· ···················· ···················· ····················
Q  8009 hCoV-19/England/MILK-ABBD12/2020|EPI_ISL_631267|20   ···················· ···················· ···················· ···················· ···················· ····················
Q  8010 hCoV-19/England/CAMC-AAF5A0/2020|EPI_ISL_631268|20   ···················· ·A·················· ···················· ···················· ···················· ··A·················
Q  8011 hCoV-19/England/ALDP-AC12A1/2020|EPI_ISL_631269|20   ···················· ···················· ···················· ···················· ···················· ····················
Q  8012 hCoV-19/England/CAMC-AAF591/2020|EPI_ISL_631270|20   ···················· ···················· ···················· ···················· ···················· ····················
Q  8013 hCoV-19/England/CAMC-AAF8D4/2020|EPI_ISL_631271|20   ···················· ···················· ···················· ···················· ···················· ····················
Q  8014 hCoV-19/England/CAMC-AAF564/2020|EPI_ISL_631272|20   ···················· ···A················ ···················· ···················· ···················· ····················
Q  8015 hCoV-19/England/CAMC-AAF607/2020|EPI_ISL_631273|20   ···················· ···················· ···················· ···················· ···················· ····················
Q  8016 hCoV-19/England/CAMC-AAF76E/2020|EPI_ISL_631274|20   ···················· ···················· ···················· ···················· ···················· ····················
Q  8017 hCoV-19/England/MILK-ABC379/2020|EPI_ISL_631275|20   ···················· ···················· ···················· ···················· ···················· ····················
Q  8018 hCoV-19/Germany/NW-UBI-23/2020|EPI_ISL_631277|2020   ···················· ···················· ···················· ···················· ···················· ····················
Q  8019 hCoV-19/Germany/NW-UBI-24/2020|EPI_ISL_631278|2020   ···················· ···················· ···················· ···················· ···················· ····················
Q  8020 hCoV-19/Germany/NW-UBI-25/2020|EPI_ISL_631279|2020   ···················· ···················· ···················· ···················· ···················· ····················
Q  8021 hCoV-19/Germany/NW-UBI-26/2020|EPI_ISL_631280|2020   ···················· ···················· ···················· ···················· ···················· ····················
Q  8022 hCoV-19/Germany/NW-UBI-27/2020|EPI_ISL_631281|2020   ···················· ···················· ···················· ···················· ···················· ····················
Q  8023 hCoV-19/Germany/NW-UBI-28/2020|EPI_ISL_631282|2020   ···················· ···················· ···················· ···················· ···················· ····················
Q  8024 hCoV-19/Germany/NW-UBI-29/2020|EPI_ISL_631283|2020   ···················· ···················· ···················· ···················· ···················· ····················
Q  8025 hCoV-19/Germany/NW-UBI-30/2020|EPI_ISL_631284|2020   ···················· ···················· ···················· ···················· ···················· ····················
Q  8026 hCoV-19/Germany/NW-UBI-31/2020|EPI_ISL_631285|2020   ···················· ···················· ···················· ···················· ···················· ····················
Q  8027 hCoV-19/Germany/NW-UBI-32/2020|EPI_ISL_631286|2020   ···················· ···················· ···················· ···················· ···················· ····················
Q  8028 hCoV-19/Germany/NW-UBI-33/2020|EPI_ISL_631287|2020   ···················· ···················· ···················· ···················· ···················· ····················
Q  8029 hCoV-19/Germany/NW-UBI-34/2020|EPI_ISL_631288|2020   ···················· ···················· ···················· ···················· ···················· ····················
Q  8030 hCoV-19/Germany/NW-UBI-35/2020|EPI_ISL_631289|2020   ···················· ···················· ···················· ···················· ···················· ····················
Q  8031 hCoV-19/Germany/NW-UBI-36/2020|EPI_ISL_631290|2020   ···················· ···················· ···················· ···················· ···················· ····················
Q  8032 hCoV-19/Germany/NW-UBI-37/2020|EPI_ISL_631291|2020   ···················· ···················· ···················· ···················· ···················· ····················
Q  8033 hCoV-19/Germany/NW-UBI-38/2020|EPI_ISL_631292|2020   ···················· ···················· ···················· ···················· ···················· ····················
Q  8034 hCoV-19/Germany/NW-UBI-39/2020|EPI_ISL_631293|2020   ···················· ···················· ···················· ···················· ···················· ····················
Q  8035 hCoV-19/Germany/NW-UBI-40/2020|EPI_ISL_631294|2020   ···················· ···················· ···················· ···················· ···················· ····················
Q  8036 hCoV-19/Germany/NW-UBI-41/2020|EPI_ISL_631295|2020   ···················· ···················· ···················· ···················· ···················· ····················
Q  8037 hCoV-19/Germany/NW-UBI-42/2020|EPI_ISL_631296|2020   ···················· ···················· ···················· ···················· ···················· ····················
Q  8038 hCoV-19/Germany/NW-UBI-43/2020|EPI_ISL_631297|2020   ···················· ···················· ···················· ···················· ···················· ····················
Q  8039 hCoV-19/Germany/NW-UBI-44/2020|EPI_ISL_631298|2020   ···················· ···················· ···················· ···················· ···················· ····················
Q  8040 hCoV-19/Germany/NW-UBI-45/2020|EPI_ISL_631299|2020   ···················· ···················· ···················· ···················· ···················· ····················
Q  8041 hCoV-19/Netherlands/UT-EMC-96/2020|EPI_ISL_632500|    ···················· ···················· ···················· ···················· ···················· ····················
Q  8042 hCoV-19/Netherlands/NH-EMC-71/2020|EPI_ISL_632501|    ···················· ···················· ···················· ···················· ···················· ····················
Q  8043 hCoV-19/Netherlands/UT-EMC-104/2020|EPI_ISL_632502    ···················· ···················· ···················· ···················· ···················· ····················
Q  8044 hCoV-19/Netherlands/ZH-EMC-662/2020|EPI_ISL_632503    ···················· ···················· ···················· ···················· ···················· ····················
Q  8045 hCoV-19/Netherlands/ZH-EMC-663/2020|EPI_ISL_632504    ···················· ···················· ···················· ···················· ···················· ····················
Q  8046 hCoV-19/Netherlands/ZH-EMC-664/2020|EPI_ISL_632505    ···················· ···················· ···················· ···················· ···················· ····················
Q  8047 hCoV-19/Netherlands/ZH-EMC-665/2020|EPI_ISL_632506    ···················· ···················· ···················· ···················· ·····T·············· ····················
Q  8048 hCoV-19/Netherlands/ZH-EMC-667/2020|EPI_ISL_632507    ···················· ···················· ···················· ···················· ···················· ····················
Q  8049 hCoV-19/Netherlands/ZH-EMC-668/2020|EPI_ISL_632508    ···················· ···················· ···················· ···················· ···················· ····················
Q  8050 hCoV-19/Netherlands/ZH-EMC-669/2020|EPI_ISL_632509    ···················· ···················· ···················· ···················· ···················· ····················
Q  8051 hCoV-19/Netherlands/NB-EMC-298/2020|EPI_ISL_632510    ···················· ···················· ···················· ···················· ···················· ····················
Q  8052 hCoV-19/Netherlands/NB-EMC-299/2020|EPI_ISL_632511    ···················· ···················· ···················· ···················· ···················· ····················
Q  8053 hCoV-19/Netherlands/ZH-EMC-670/2020|EPI_ISL_632512    ···················· ···················· ···················· ···················· ···················· ····················
```

| | | | | | | | |
|---|---|---|---|---|---|---|---|
| Q | 8054 hCoV-19/Netherlands/ZH-EMC-671/2020\|EPI_ISL_632513 | ···················· | ···················· | ····················· | ···················· | ···················· | ···················· |
| Q | 8055 hCoV-19/Netherlands/ZH-EMC-673/2020\|EPI_ISL_632514 | ···················· | ···················· | ····················· | ···················· | ···················· | ···················· |
| Q | 8056 hCoV-19/Netherlands/ZH-EMC-674/2020\|EPI_ISL_632515 | ···················· | ···················· | ····················· | ···················· | ···················· | ···················· |
| Q | 8057 hCoV-19/Netherlands/ZH-EMC-676/2020\|EPI_ISL_632516 | ···················· | ···················· | ····················· | ···················· | ···················· | ···················· |
| Q | 8058 hCoV-19/Netherlands/ZH-EMC-677/2020\|EPI_ISL_632517 | ···················· | ···················· | ····················· | ···················· | ···················· | ···················· |
| Q | 8059 hCoV-19/Netherlands/ZH-EMC-678/2020\|EPI_ISL_632518 | ···················· | ·A·················· | ····················· | ···················· | ···················· | ··A················· |
| Q | 8060 hCoV-19/Netherlands/ZH-EMC-679/2020\|EPI_ISL_632519 | ···················· | ···················· | ····················· | ···················· | ···················· | ···················· |
| Q | 8061 hCoV-19/Netherlands/NH-EMC-72/2020\|EPI_ISL_632520\| | ···················· | ···················· | ····················· | ···················· | ···················· | ···················· |
| Q | 8062 hCoV-19/Netherlands/NH-EMC-73/2020\|EPI_ISL_632521\| | ···················· | ···················· | ····················· | ···················· | ···················· | ···················· |
| Q | 8063 hCoV-19/Netherlands/NH-EMC-74/2020\|EPI_ISL_632522\| | ···················· | ···················· | ····················· | ···················· | ···················· | ···················· |
| Q | 8064 hCoV-19/Netherlands/NH-EMC-75/2020\|EPI_ISL_632523\| | ···················· | ···················· | ····················· | ···················· | ···················· | ···················· |
| Q | 8065 hCoV-19/Netherlands/NH-EMC-76/2020\|EPI_ISL_632524\| | ···················· | ···················· | ····················· | ···················· | ···················· | ···················· |
| Q | 8066 hCoV-19/Netherlands/NH-EMC-78/2020\|EPI_ISL_632525\| | ···················· | ···················· | ····················· | ···················· | ···················· | ···················· |
| Q | 8067 hCoV-19/Netherlands/NH-EMC-79/2020\|EPI_ISL_632526\| | ···················· | ···················· | ····················· | ···················· | ···················· | ···················· |
| Q | 8068 hCoV-19/Netherlands/NH-EMC-82/2020\|EPI_ISL_632527\| | ···················· | ···················· | ····················· | ···················· | ···················· | ···················· |
| Q | 8069 hCoV-19/Netherlands/NH-EMC-81/2020\|EPI_ISL_632528\| | ···················· | ···················· | ····················· | ···················· | ···················· | ···················· |
| Q | 8070 hCoV-19/Netherlands/NH-EMC-83/2020\|EPI_ISL_632529\| | ···················· | ···················· | ····················· | ···················· | ···················· | ···················· |
| Q | 8071 hCoV-19/Netherlands/ZH-EMC-681/2020\|EPI_ISL_632530 | ···················· | ···················· | ····················· | ···················· | ······T·······G····· | ···················· |
| Q | 8072 hCoV-19/Netherlands/UT-EMC-106/2020\|EPI_ISL_632531 | ···················· | ···················· | ····················· | ···················· | ······T············· | ···················· |
| Q | 8073 hCoV-19/Netherlands/NH-EMC-85/2020\|EPI_ISL_632532\| | ···················· | ···················· | ····················· | ···················· | ···················· | ···················· |
| Q | 8074 hCoV-19/Netherlands/NH-EMC-86/2020\|EPI_ISL_632533\| | ···················· | ···················· | ····················· | ···················· | ···················· | ···················· |
| Q | 8075 hCoV-19/Netherlands/NH-EMC-88/2020\|EPI_ISL_632534\| | ···················· | ···················· | ····················· | ···················· | ···················· | ···················· |
| Q | 8076 hCoV-19/Netherlands/NB-EMC-300/2020\|EPI_ISL_632535 | ···················· | ···················· | ····················· | ···················· | ···················· | ···················· |
| Q | 8077 hCoV-19/Netherlands/NB-EMC-301/2020\|EPI_ISL_632536 | ···················· | ···················· | ····················· | ···················· | ···················· | ···················· |
| Q | 8078 hCoV-19/Netherlands/NB-EMC-302/2020\|EPI_ISL_632537 | ···················· | ···················· | ····················· | ···················· | ···················· | ···················· |
| Q | 8079 hCoV-19/Netherlands/NB-EMC-304/2020\|EPI_ISL_632538 | ···················· | ···················· | ····················· | ···················· | ···················· | ···················· |
| Q | 8080 hCoV-19/Netherlands/ZH-EMC-683/2020\|EPI_ISL_632539 | ···················· | ···················· | ····················· | ···················· | ···················· | ···················· |
| Q | 8081 hCoV-19/Netherlands/ZH-EMC-685/2020\|EPI_ISL_632540 | ···················· | ···················· | ····················· | ···················· | ···················· | ···················· |
| Q | 8082 hCoV-19/Netherlands/ZH-EMC-686/2020\|EPI_ISL_632541 | ···················· | ···················· | ····················· | ···················· | ···················· | ···················· |
| Q | 8083 hCoV-19/Netherlands/ZH-EMC-689/2020\|EPI_ISL_632543 | ···················· | ···················· | ····················· | ···················· | ···················· | ···················· |
| Q | 8084 hCoV-19/Netherlands/ZH-EMC-710/2020\|EPI_ISL_632557 | ···················· | ···················· | ····················· | ···················· | ···················· | ···················· |
| Q | 8085 hCoV-19/Netherlands/ZH-EMC-711/2020\|EPI_ISL_632558 | ···················· | ···················· | ····················· | ···················· | ···················· | ···················· |
| Q | 8086 hCoV-19/Netherlands/ZH-EMC-712/2020\|EPI_ISL_632559 | ···················· | ···················· | ····················· | ···················· | ···················· | ···················· |
| Q | 8087 hCoV-19/Netherlands/ZH-EMC-713/2020\|EPI_ISL_632560 | ···················· | ···················· | ····················· | ···················· | ···················· | ···················· |
| Q | 8088 hCoV-19/Netherlands/ZH-EMC-714/2020\|EPI_ISL_632561 | ···················· | ···················· | ····················· | ···················· | ···················· | ···················· |
| Q | 8089 hCoV-19/Netherlands/ZH-EMC-715/2020\|EPI_ISL_632562 | ···················· | ···················· | ····················· | ···················· | ···················· | ···················· |
| Q | 8090 hCoV-19/Netherlands/ZH-EMC-716/2020\|EPI_ISL_632563 | ···················· | ···················· | ····················· | ···················· | ···················· | ···················· |
| Q | 8091 hCoV-19/Netherlands/ZH-EMC-717/2020\|EPI_ISL_632564 | ···················· | ···················· | ····················· | ···················· | ···················· | ···················· |
| Q | 8092 hCoV-19/Netherlands/ZH-EMC-718/2020\|EPI_ISL_632565 | ···················· | ···················· | ····················· | ···················· | ···················· | ···················· |
| Q | 8093 hCoV-19/Netherlands/ZH-EMC-719/2020\|EPI_ISL_632566 | ···················· | ···················· | ····················· | ···················· | ···················· | ···················· |
| Q | 8094 hCoV-19/Netherlands/ZH-EMC-720/2020\|EPI_ISL_632567 | ···················· | ···················· | ····················· | ···················· | ······T············· | ···················· |
| Q | 8095 hCoV-19/Netherlands/ZH-EMC-721/2020\|EPI_ISL_632568 | ···················· | ···················· | ····················· | ···················· | ···················· | ···················· |
| Q | 8096 hCoV-19/Netherlands/ZH-EMC-722/2020\|EPI_ISL_632569 | ···················· | ···················· | ····················· | ···················· | ······T············· | ···················· |
| Q | 8097 hCoV-19/Netherlands/ZH-EMC-723/2020\|EPI_ISL_632570 | ···················· | ···A················ | ····················· | ···················· | ···················· | ···················· |
| Q | 8098 hCoV-19/Netherlands/ZH-EMC-724/2020\|EPI_ISL_632571 | ···················· | ···················· | ····················· | ···················· | ······T············· | ···················· |
| Q | 8099 hCoV-19/Netherlands/ZH-EMC-726/2020\|EPI_ISL_632572 | ···················· | ···················· | ····················· | ···················· | ···················· | ···················· |
| Q | 8100 hCoV-19/Netherlands/ZH-EMC-730/2020\|EPI_ISL_632573 | ···················· | ···················· | ····················· | ···················· | ···················· | ···················· |
| Q | 8101 hCoV-19/Netherlands/ZH-EMC-731/2020\|EPI_ISL_632574 | ···················· | ···················· | ····················· | ···················· | ···················· | ···················· |
| Q | 8102 hCoV-19/Netherlands/ZH-EMC-732/2020\|EPI_ISL_632575 | ···················· | ···················· | ····················· | ···················· | ···················· | ···················· |
| Q | 8103 hCoV-19/Netherlands/ZH-EMC-733/2020\|EPI_ISL_632576 | ···················· | ···················· | ····················· | ···················· | ···················· | ···················· |
| Q | 8104 hCoV-19/Netherlands/ZH-EMC-734/2020\|EPI_ISL_632577 | ···················· | ···················· | ····················· | ···················· | ···················· | ···················· |
| Q | 8105 hCoV-19/Netherlands/ZH-EMC-736/2020\|EPI_ISL_632578 | ···················· | ···················· | ····················· | ···················· | ···················· | ···················· |
| Q | 8106 hCoV-19/Netherlands/ZH-EMC-737/2020\|EPI_ISL_632579 | ···················· | ···················· | ····················· | ···················· | ···················· | ···················· |
| Q | 8107 hCoV-19/Netherlands/ZH-EMC-738/2020\|EPI_ISL_632580 | ···················· | ···················· | ····················· | ···················· | ···················· | ···················· |
| Q | 8108 hCoV-19/Netherlands/ZH-EMC-739/2020\|EPI_ISL_632581 | ···················· | ···················· | ····················· | ···················· | ···················· | ···················· |
| Q | 8109 hCoV-19/Netherlands/ZH-EMC-740/2020\|EPI_ISL_632582 | ···················· | ···················· | ····················· | ···················· | ···················· | ···················· |
| Q | 8110 hCoV-19/Netherlands/ZH-EMC-742/2020\|EPI_ISL_632583 | ···················· | ···················· | ····················· | ···················· | ···················· | ···················· |
| Q | 8111 hCoV-19/Netherlands/ZH-EMC-743/2020\|EPI_ISL_632584 | ···················· | ···················· | ····················· | ···················· | ···················· | ···················· |
| Q | 8112 hCoV-19/Netherlands/ZH-EMC-744/2020\|EPI_ISL_632585 | ···················· | ···················· | ····················· | ···················· | ···················· | ···················· |
| Q | 8113 hCoV-19/Netherlands/ZH-EMC-745/2020\|EPI_ISL_632586 | ···················· | ···················· | ····················· | ···················· | ···················· | ···················· |
| Q | 8114 hCoV-19/Netherlands/ZH-EMC-746/2020\|EPI_ISL_632587 | ···················· | ···················· | ····················· | ···················· | ···················· | ···················· |
| Q | 8115 hCoV-19/Netherlands/ZH-EMC-747/2020\|EPI_ISL_632588 | ···················· | ···················· | ····················· | ···················· | ···················· | ···················· |
| Q | 8116 hCoV-19/Netherlands/ZH-EMC-748/2020\|EPI_ISL_632589 | ···················· | ···················· | ····················· | ···················· | ···················· | ···················· |
| Q | 8117 hCoV-19/Netherlands/ZH-EMC-749/2020\|EPI_ISL_632590 | ···················· | ···················· | ····················· | ···················· | ···················· | ···················· |
| Q | 8118 hCoV-19/Netherlands/ZH-EMC-750/2020\|EPI_ISL_632591 | ···················· | ···················· | ····················· | ···················· | ···················· | ···················· |
| Q | 8119 hCoV-19/Netherlands/ZH-EMC-751/2020\|EPI_ISL_632592 | ···················· | ···A················ | ····················· | ···················· | ···················· | ···················· |
| Q | 8120 hCoV-19/Netherlands/ZH-EMC-752/2020\|EPI_ISL_632593 | ···················· | ···················· | ····················· | ···················· | ···················· | ···················· |
| Q | 8121 hCoV-19/Netherlands/ZH-EMC-753/2020\|EPI_ISL_632594 | ···················· | ···················· | ····················· | ···················· | ···················· | ···················· |
| Q | 8122 hCoV-19/Netherlands/ZH-EMC-754/2020\|EPI_ISL_632595 | ···················· | ···················· | ····················· | ···················· | ···················· | ···················· |
| Q | 8123 hCoV-19/Netherlands/ZH-EMC-759/2020\|EPI_ISL_632596 | ···················· | ···················· | ····················· | ···················· | ···················· | ···················· |
| Q | 8124 hCoV-19/Netherlands/ZH-EMC-760/2020\|EPI_ISL_632597 | ···················· | ···················· | ····················· | ···················· | ···················· | ···················· |
| Q | 8125 hCoV-19/Netherlands/ZH-EMC-761/2020\|EPI_ISL_632598 | ···················· | ···A················ | ····················· | ···················· | ···················· | ···················· |
| Q | 8126 hCoV-19/Netherlands/ZH-EMC-762/2020\|EPI_ISL_632599 | ···················· | ···A················ | ····················· | ···················· | ···················· | ···················· |
| Q | 8127 hCoV-19/England/QEUH-AD2850/2020\|EPI_ISL_633822\|20 | ···················· | ···················· | ····················· | ···················· | ···················· | ···················· |
| Q | 8128 hCoV-19/England/QEUH-AD5DA9/2020\|EPI_ISL_633823\|20 | ···················· | ···················· | ····················· | ···················· | ···················· | ···················· |
| Q | 8129 hCoV-19/England/QEUH-AD14E4/2020\|EPI_ISL_633824\|20 | ···················· | ·A·················· | ····················· | ···················· | ···················· | ···················· |
| Q | 8130 hCoV-19/England/QEUH-AD2805/2020\|EPI_ISL_633826\|20 | ···················· | ·A·················· | ····················· | ···················· | ···················· | ··A················· |
| Q | 8131 hCoV-19/England/QEUH-AD0DCC/2020\|EPI_ISL_633827\|20 | ···················· | ···················· | ····················· | ···················· | ···················· | ···················· |
| Q | 8132 hCoV-19/England/QEUH-AD3303/2020\|EPI_ISL_633829\|20 | ···················· | ···················· | ····················· | ···················· | ···················· | ···················· |
| Q | 8133 hCoV-19/England/QEUH-AD0A01/2020\|EPI_ISL_633830\|20 | ···················· | ···················· | ····················· | ···················· | ···················· | ···················· |

```
Q 8134 hCoV-19/England/QEUH-AD289C/2020|EPI_ISL_633832|20 ···················· ···················· ···················· ···················· ···················· ····················
Q 8135 hCoV-19/England/QEUH-AD59E1/2020|EPI_ISL_633834|20 ···················· ···················· ···················· ···················· ···················· ····················
Q 8136 hCoV-19/England/QEUH-AD0852/2020|EPI_ISL_633836|20 ···················· ···················· ···················· ···················· ···················· ····················
Q 8137 hCoV-19/England/QEUH-AD10B3/2020|EPI_ISL_633837|20 ···················· ···················· ···················· ···················· ···················· ····················
Q 8138 hCoV-19/England/QEUH-AD0DAE/2020|EPI_ISL_633838|20 ···················· ···················· ···················· ···················· ·····T·············· ····················
Q 8139 hCoV-19/England/QEUH-AD1BEF/2020|EPI_ISL_633839|20 ···················· ···················· ···················· ···················· ···················· ····················
Q 8140 hCoV-19/England/QEUH-AD5608/2020|EPI_ISL_633840|20 ···················· ···················· ···················· ···················· ···················· ····················
Q 8141 hCoV-19/England/QEUH-AD15E1/2020|EPI_ISL_633841|20 ···················· ···················· ···················· ···················· ···················· ····················
Q 8142 hCoV-19/England/QEUH-AD4E7A/2020|EPI_ISL_633842|20 ···················· ···················· ···················· ···················· ···················· ····················
Q 8143 hCoV-19/England/QEUH-AD5D03/2020|EPI_ISL_633843|20 ···················· ···················· ···················· ···················· ···················· ····················
Q 8144 hCoV-19/England/QEUH-AD38AA/2020|EPI_ISL_633844|20 ···················· ···················· ···················· ···················· ···················· ····················
Q 8145 hCoV-19/England/QEUH-AD54A4/2020|EPI_ISL_633846|20 ···················· ···················· ···················· ···················· ···················· ····················
Q 8146 hCoV-19/England/QEUH-AD1BA3/2020|EPI_ISL_633847|20 ···················· ···················· ···················· ···················· ···················· ····················
Q 8147 hCoV-19/England/QEUH-AD0C0B/2020|EPI_ISL_633848|20 ···················· ···················· ···················· ···················· ···················· ····················
Q 8148 hCoV-19/England/QEUH-AD12EA/2020|EPI_ISL_633850|20 ···················· ·A·················· ···················· ···················· ···················· ···A················
Q 8149 hCoV-19/England/QEUH-AD38F5/2020|EPI_ISL_633852|20 ···················· ···················· ···················· ···················· ···················· ····················
Q 8150 hCoV-19/England/QEUH-AD0843/2020|EPI_ISL_633853|20 ···················· ···················· ···················· ···················· ···················· ····················
Q 8151 hCoV-19/England/QEUH-AD4FFF/2020|EPI_ISL_633854|20 ···················· ···················· ···················· ···················· ···················· ····················
Q 8152 hCoV-19/England/QEUH-AD1314/2020|EPI_ISL_633855|20 ···················· ···················· ···················· ···················· ···················· ····················
Q 8153 hCoV-19/England/QEUH-AD3619/2020|EPI_ISL_633856|20 ···················· ···················· ···················· ···················· ···················· ····················
Q 8154 hCoV-19/England/QEUH-AD3558/2020|EPI_ISL_633857|20 ···················· ···················· ···················· ···················· ···················· ····················
Q 8155 hCoV-19/England/QEUH-AD3901/2020|EPI_ISL_633858|20 ···················· ···················· ···················· ···················· ···················· ····················
Q 8156 hCoV-19/England/QEUH-AD5DF4/2020|EPI_ISL_633859|20 ···················· ···················· ···················· ···················· ···················· ····················
Q 8157 hCoV-19/England/QEUH-AD392F/2020|EPI_ISL_633861|20 ···················· ···················· ···················· ···················· ···················· ····················
Q 8158 hCoV-19/England/QEUH-AD54D1/2020|EPI_ISL_633862|20 ···················· ···················· ···················· ···················· ···················· ····················
Q 8159 hCoV-19/England/QEUH-AD55A1/2020|EPI_ISL_633863|20 ···················· ···················· ···················· ···················· ···················· ····················
Q 8160 hCoV-19/England/QEUH-AD4F3B/2020|EPI_ISL_633864|20 ···················· ···················· ···················· ···················· ···················· ····················
Q 8161 hCoV-19/England/QEUH-AD15B4/2020|EPI_ISL_633865|20 ···················· ·A·················· ···················· ···················· ···················· ····················
Q 8162 hCoV-19/England/QEUH-AD0816/2020|EPI_ISL_633866|20 ···················· ···················· ···················· ···················· ···················· ····················
Q 8163 hCoV-19/England/QEUH-AD1B58/2020|EPI_ISL_633867|20 ···················· ···················· ···················· ···················· ···················· ····················
Q 8164 hCoV-19/England/QEUH-AD59B4/2020|EPI_ISL_633868|20 ···················· ···················· ···················· ···················· ···················· ····················
Q 8165 hCoV-19/England/QEUH-AD1183/2020|EPI_ISL_633869|20 ···················· ···················· ···················· ···················· ···················· ····················
Q 8166 hCoV-19/England/QEUH-AD0834/2020|EPI_ISL_633870|20 ···················· ···················· ···················· ···················· ···················· ····················
Q 8167 hCoV-19/England/QEUH-AD4FE0/2020|EPI_ISL_633871|20 ···················· ···················· ···················· ···················· ···················· ····················
Q 8168 hCoV-19/England/QEUH-AD352B/2020|EPI_ISL_633874|20 ···················· ···················· ···················· ···················· ···················· ····················
Q 8169 hCoV-19/England/QEUH-AD0E41/2020|EPI_ISL_633888|20 ···················· ···················· ···················· ···················· ···················· ····················
Q 8170 hCoV-19/England/QEUH-AD396B/2020|EPI_ISL_633890|20 ···················· ···················· ···················· ···················· ···················· ····················
Q 8171 hCoV-19/England/QEUH-AD161B/2020|EPI_ISL_633891|20 ···················· ···················· ···················· ···················· ···················· ····················
Q 8172 hCoV-19/England/QEUH-AD279F/2020|EPI_ISL_633892|20 ···················· ···················· ···················· ···················· ···················· ····················
Q 8173 hCoV-19/England/QEUH-AD31FA/2020|EPI_ISL_633894|20 ···················· ···················· ···················· ···················· ···················· ····················
Q 8174 hCoV-19/England/QEUH-AD4E2F/2020|EPI_ISL_633895|20 ···················· ···················· ···················· ···················· ···················· ····················
Q 8175 hCoV-19/England/QEUH-AD08CB/2020|EPI_ISL_633896|20 ···················· ···················· ···················· ···················· ···················· ····················
Q 8176 hCoV-19/England/QEUH-AD5D5E/2020|EPI_ISL_633897|20 ···················· ···················· ···················· ···················· ···················· ····················
Q 8177 hCoV-19/England/QEUH-AD3910/2020|EPI_ISL_633898|20 ···················· ···················· ···················· ···················· ···················· ····················
Q 8178 hCoV-19/England/QEUH-AD0DDB/2020|EPI_ISL_633899|20 ···················· ···················· ···················· ···················· ···················· ····················
Q 8179 hCoV-19/USA/CA-ALSR-3394/2020|EPI_ISL_636001|2020- ···················· ·A·················· ···················· ···················· ···················· ····················
Q 8180 hCoV-19/USA/CA-ALSR-3396/2020|EPI_ISL_636002|2020- ···················· ···················· ···················· ···················· ···················· ····················
Q 8181 hCoV-19/USA/CA-ALSR-3399/2020|EPI_ISL_636003|2020- ···················· ·A·················· ···················· ···················· ···················· ····················
Q 8182 hCoV-19/USA/CA-ALSR-3400/2020|EPI_ISL_636004|2020- ···················· ···················· ···················· ···················· ···················· ····················
Q 8183 hCoV-19/USA/CA-ALSR-3402/2020|EPI_ISL_636005|2020- ···················· ·A·················· ···················· ···················· ···················· ····················
Q 8184 hCoV-19/USA/CA-ALSR-3404/2020|EPI_ISL_636006|2020- ···················· ·A·················· ···················· ···················· ···················· ····················
Q 8185 hCoV-19/USA/CA-ALSR-3408/2020|EPI_ISL_636010|2020- ···················· ·A·················· ···················· ···················· ···················· ····················
Q 8186 hCoV-19/USA/CA-ALSR-3409/2020|EPI_ISL_636011|2020- ···················· ·A·················· ···················· ···················· ···················· ····················
Q 8187 hCoV-19/USA/CA-ALSR-3410/2020|EPI_ISL_636012|2020- ···················· ·A·················· ···················· ···················· ···················· ····················
Q 8188 hCoV-19/USA/CA-ALSR-3413/2020|EPI_ISL_636014|2020- ···················· ·A·················· ···················· ···················· ···················· ····················
Q 8189 hCoV-19/USA/CA-ALSR-3418/2020|EPI_ISL_636016|2020- ···················· ·A·················· ···················· ···················· ···················· ····················
Q 8190 hCoV-19/USA/CA-ALSR-3419/2020|EPI_ISL_636017|2020- ···················· ·A·················· ···················· ···················· ···················· ····················
Q 8191 hCoV-19/USA/CA-ALSR-3420/2020|EPI_ISL_636018|2020- ···················· ···················· ···················· ···················· ···················· ····················
Q 8192 hCoV-19/USA/CA-ALSR-3422/2020|EPI_ISL_636019|2020- ···················· ·A·················· ···················· ···················· ···················· ····················
Q 8193 hCoV-19/Czech Republic/NRL_9888/2020|EPI_ISL_62661 ···················· ···················· ···················· ···················· ···················· ····················
Q 8194 hCoV-19/Czech Republic/NRL_9889/2020|EPI_ISL_62661 ···················· ···················· ···················· ···················· ···················· ····················
Q 8195 hCoV-19/Czech Republic/NRL_9891/2020|EPI_ISL_62661 ···················· ···················· ···················· ···················· ···················· ····················
Q 8196 hCoV-19/Czech Republic/NRL_9893/2020|EPI_ISL_62662 ···················· ···················· ···················· ···················· ···················· ····················
Q 8197 hCoV-19/Czech Republic/NRL_9894/2020|EPI_ISL_62662 ···················· ···················· ···················· ···················· ···················· ····················
Q 8198 hCoV-19/Czech Republic/NRL_9899/2020|EPI_ISL_62662 ···················· ···················· ···················· ···················· ···················· ····················
Q 8199 hCoV-19/Czech Republic/NRL_9903/2020|EPI_ISL_62662 ···················· ···················· ···················· ···················· ···················· ····················
Q 8200 hCoV-19/Czech Republic/NRL_10178/2020|EPI_ISL_6266 ···················· ···················· ···················· ···················· ···················· ····················
Q 8201 hCoV-19/Czech Republic/NRL_10181/2020|EPI_ISL_6266 ···················· ···················· ···················· ···················· ···················· ····················
Q 8202 hCoV-19/Singapore/1119/2020|EPI_ISL_626630|2020-10 ···················· ···················· ···················· ···················· ···················· ····················
Q 8203 hCoV-19/Singapore/1124/2020|EPI_ISL_626635|2020-10 ···················· ···················· ···················· ···················· ···················· ····················
Q 8204 hCoV-19/Singapore/1127/2020|EPI_ISL_626639|2020-10 ···················· ···················· ···················· ···················· ···················· ····················
Q 8205 hCoV-19/Singapore/1128/2020|EPI_ISL_626640|2020-10 ···················· ·A·················· ···················· ···················· ···················· ···A················
Q 8206 hCoV-19/Singapore/1131/2020|EPI_ISL_626641|2020-10 ···················· ···················· ···················· ···················· ···················· ····················
Q 8207 hCoV-19/Singapore/1130/2020|EPI_ISL_626642|2020-10 ···················· ···················· ···················· ···················· ···················· ···A················
Q 8208 hCoV-19/Singapore/1132/2020|EPI_ISL_626643|2020-10 ···················· ···················· ···················· ···················· ···················· ····················
Q 8209 hCoV-19/Wales/PHWC-483F6E/2020|EPI_ISL_627948|2020 ···················· ···················· ···················· ···················· ···················· ····················
Q 8210 hCoV-19/Wales/PHWC-483F7D/2020|EPI_ISL_627949|2020 ···················· ···················· ···················· ···················· ···················· ····················
Q 8211 hCoV-19/Wales/PHWC-483F8C/2020|EPI_ISL_627950|2020 ···················· ···················· ···················· ···················· ···················· ····················
Q 8212 hCoV-19/Wales/PHWC-483FAA/2020|EPI_ISL_627951|2020 ···················· ···················· ···················· ···················· ···················· ····················
Q 8213 hCoV-19/Wales/PHWC-483FC8/2020|EPI_ISL_627952|2020 ···················· ···················· ···················· ···················· ···················· ····················
```

| | | | | | | | | |
|---|---|---|---|---|---|---|---|---|
| Q | 8214 hCoV-19/Wales/PHWC-483FE6/2020\|EPI_ISL_627953\|2020 | ···················· | ···················· | ···················· | ···················· | ···················· | ···················· | |
| Q | 8215 hCoV-19/Wales/PHWC-483FF5/2020\|EPI_ISL_627954\|2020 | ···················· | ···················· | ···················· | ···················· | ···················· | ···················· | |
| Q | 8216 hCoV-19/Wales/PHWC-484000/2020\|EPI_ISL_627955\|2020 | ···················· | ···················· | ···················· | ···················· | ···················· | ···················· | |
| Q | 8217 hCoV-19/Wales/PHWC-48401F/2020\|EPI_ISL_627956\|2020 | ···················· | ···················· | ···················· | ···················· | ···················· | ···················· | |
| Q | 8218 hCoV-19/Wales/PHWC-48402E/2020\|EPI_ISL_627957\|2020 | ···················· | ···················· | ···················· | ···················· | ···················· | ···················· | |
| Q | 8219 hCoV-19/Wales/PHWC-48403D/2020\|EPI_ISL_627958\|2020 | ···················· | ···················· | ···················· | ···················· | ···················· | ···················· | |
| Q | 8220 hCoV-19/Wales/PHWC-48406A/2020\|EPI_ISL_627960\|2020 | ···················· | ···················· | ···················· | ···················· | ···················· | ···················· | |
| Q | 8221 hCoV-19/Wales/PHWC-484079/2020\|EPI_ISL_627961\|2020 | ···················· | ···················· | ···················· | ···················· | ···················· | ···················· | |
| Q | 8223 hCoV-19/Wales/PHWC-48410D/2020\|EPI_ISL_627967\|2020 | ···················· | ···················· | ···················· | ···················· | ···················· | ···················· | |
| Q | 8225 hCoV-19/Wales/PHWC-484149/2020\|EPI_ISL_627970\|2020 | ···················· | ···················· | ···················· | ···················· | ···················· | ···················· | |
| Q | 8226 hCoV-19/Wales/PHWC-484167/2020\|EPI_ISL_627971\|2020 | ···················· | ···················· | ···················· | ···················· | ···················· | ···················· | |
| Q | 8227 hCoV-19/Wales/PHWC-484185/2020\|EPI_ISL_627972\|2020 | ···················· | ···················· | ···················· | ···················· | ···················· | ···················· | |
| Q | 8228 hCoV-19/Wales/PHWC-4841A3/2020\|EPI_ISL_627973\|2020 | ···················· | ···················· | ···················· | ···················· | ···················· | ···················· | |
| Q | 8229 hCoV-19/Wales/PHWC-4841B2/2020\|EPI_ISL_627974\|2020 | ···················· | ···················· | ···················· | ···················· | ···················· | ···················· | |
| Q | 8230 hCoV-19/Wales/PHWC-4841C1/2020\|EPI_ISL_627975\|2020 | ···················· | ···················· | ···················· | ···················· | ···················· | ···················· | |
| Q | 8231 hCoV-19/Wales/PHWC-4841EF/2020\|EPI_ISL_627976\|2020 | ···················· | ···················· | ···················· | ···················· | ···················· | ···················· | |
| Q | 8232 hCoV-19/Wales/PHWC-48420A/2020\|EPI_ISL_627977\|2020 | ···················· | ···················· | ···················· | ···················· | ···················· | ···················· | |
| Q | 8233 hCoV-19/Wales/PHWC-484219/2020\|EPI_ISL_627978\|2020 | ···················· | ···················· | ···················· | ···················· | ···················· | ···················· | |
| Q | 8234 hCoV-19/Wales/PHWC-484228/2020\|EPI_ISL_627979\|2020 | ···················· | ···················· | ···················· | ···················· | NNN················· | ···················· | 1 |
| Q | 8235 hCoV-19/Wales/PHWC-484237/2020\|EPI_ISL_627980\|2020 | ···················· | ···················· | ···················· | ···················· | ···················· | ···················· | |
| Q | 8236 hCoV-19/Wales/PHWC-484255/2020\|EPI_ISL_627981\|2020 | ···················· | ···················· | ···················· | ···················· | ···················· | ···················· | |
| Q | 8237 hCoV-19/Wales/PHWC-484291/2020\|EPI_ISL_627982\|2020 | ···················· | ···················· | ···················· | ···················· | ···················· | ···················· | |
| Q | 8238 hCoV-19/Wales/PHWC-4842A0/2020\|EPI_ISL_627983\|2020 | ···················· | ···················· | ···················· | ···················· | ···················· | ···················· | |
| Q | 8239 hCoV-19/Wales/PHWC-4842BF/2020\|EPI_ISL_627984\|2020 | ···················· | ···················· | ···················· | ···················· | ···················· | ···················· | |
| Q | 8240 hCoV-19/Wales/PHWC-4842DD/2020\|EPI_ISL_627985\|2020 | ···················· | ···················· | ···················· | ···················· | ···················· | ···················· | |
| Q | 8242 hCoV-19/Wales/PHWC-4842FB/2020\|EPI_ISL_627987\|2020 | ···················· | ···················· | ···················· | ···················· | ···················· | ···················· | |
| Q | 8243 hCoV-19/Wales/PHWC-484307/2020\|EPI_ISL_627988\|2020 | ···················· | ···················· | ···················· | ···················· | ···················· | ···················· | |
| Q | 8244 hCoV-19/Wales/PHWC-484316/2020\|EPI_ISL_627989\|2020 | ···················· | ···················· | ···················· | ···················· | ···················· | ···················· | |
| Q | 8245 hCoV-19/England/MILK-ABB547/2020\|EPI_ISL_631100\|20 | ···················· | ···················· | ···················· | ···················· | ···················· | ···················· | |
| Q | 8246 hCoV-19/England/CAMC-AAF5FB/2020\|EPI_ISL_631101\|20 | ···················· | ···················· | ···················· | ···················· | ···················· | ···················· | |
| Q | 8247 hCoV-19/England/MILK-ABC4B2/2020\|EPI_ISL_631103\|20 | ···················· | ···················· | ···················· | ···················· | ···················· | ···················· | |
| Q | 8248 hCoV-19/England/MILK-ABB87B/2020\|EPI_ISL_631104\|20 | ···················· | ···················· | ···················· | ···················· | ·····T·············· | ···················· | |
| Q | 8249 hCoV-19/England/CAMC-AAF731/2020\|EPI_ISL_631105\|20 | ···················· | ···················· | ···················· | ···················· | ···················· | ···················· | |
| Q | 8250 hCoV-19/England/MILK-ABC054/2020\|EPI_ISL_631106\|20 | ···················· | ···················· | ···················· | ···················· | ···················· | ···················· | |
| Q | 8251 hCoV-19/England/ALDP-AC5D60/2020\|EPI_ISL_631107\|20 | ···················· | ···················· | ···················· | ···················· | ···················· | ···················· | |
| Q | 8252 hCoV-19/England/MILK-ABBADF/2020\|EPI_ISL_631108\|20 | ···················· | ···················· | ···················· | ···················· | ·····T·············· | ···············A···· | |
| Q | 8253 hCoV-19/England/CAMC-AAF8E3/2020\|EPI_ISL_631109\|20 | ···················· | ···················· | ···················· | ···················· | ···················· | ···················· | |
| Q | 8254 hCoV-19/England/MILK-ABBFC1/2020\|EPI_ISL_631110\|20 | ···················· | ·A·······A·········· | ···················· | ···················· | ···················· | ···················· | |
| Q | 8255 hCoV-19/England/ALDP-ABD30F/2020\|EPI_ISL_631111\|20 | ···················· | ···················· | ···················· | ···················· | ···················· | ···················· | |
| Q | 8256 hCoV-19/England/ALDP-AC1274/2020\|EPI_ISL_631113\|20 | ···················· | ·A·······A·········· | ···················· | ···················· | ···················· | ···················· | |
| Q | 8257 hCoV-19/England/MILK-ABBB54/2020\|EPI_ISL_631114\|20 | ···················· | ···················· | ···················· | ···················· | ·····T·············· | ···················· | |
| Q | 8258 hCoV-19/England/MILK-ABBCBB/2020\|EPI_ISL_631115\|20 | ···················· | ···················· | ···················· | ···················· | ···················· | ···················· | |
| Q | 8259 hCoV-19/England/ALDP-ABD23F/2020\|EPI_ISL_631116\|20 | ···················· | ···················· | ···················· | ···················· | ···················· | ···················· | |
| Q | 8260 hCoV-19/England/CAMC-AAF6BC/2020\|EPI_ISL_631117\|20 | ···················· | ···················· | ···················· | ···················· | ···················· | ···················· | |
| Q | 8261 hCoV-19/England/MILK-ABBD5E/2020\|EPI_ISL_631119\|20 | ···················· | ···················· | ···················· | ···················· | ···················· | ···················· | |
| Q | 8262 hCoV-19/England/MILK-ABC458/2020\|EPI_ISL_631120\|20 | ···················· | ···················· | ···················· | ···················· | ···················· | ···················· | |
| Q | 8263 hCoV-19/England/MILK-ABC4A3/2020\|EPI_ISL_631121\|20 | ···················· | ···················· | ···················· | ···················· | ···················· | ···················· | |
| Q | 8264 hCoV-19/England/MILK-ABBF85/2020\|EPI_ISL_631122\|20 | ···················· | ···················· | ···················· | ···················· | ···················· | ···················· | |
| Q | 8265 hCoV-19/England/CAMC-AAF670/2020\|EPI_ISL_631123\|20 | ···················· | ···················· | ···················· | ···················· | ···················· | ···················· | |
| Q | 8266 hCoV-19/England/MILK-ABC3C4/2020\|EPI_ISL_631124\|20 | ···················· | ···················· | ···················· | ···················· | ·····T·············· | ···················· | |
| Q | 8267 hCoV-19/England/MILK-ABBB63/2020\|EPI_ISL_631125\|20 | ···················· | ·A·················· | ···················· | ···················· | ···················· | ··A················· | |
| Q | 8268 hCoV-19/England/MILK-ABBE97/2020\|EPI_ISL_631126\|20 | ···················· | ···················· | ···················· | ···················· | ···················· | ···················· | |
| Q | 8269 hCoV-19/mink/Netherlands/NB-EMC-55-6/2020\|EPI_ISL_ | ···················· | ···················· | ···················· | ···················· | ·····T·············· | ···················· | |
| Q | 8270 hCoV-19/mink/Netherlands/NB-EMC-55-18/2020\|EPI_ISL | ···················· | ···················· | ···················· | ···················· | ·····T·············· | ···················· | |
| Q | 8271 hCoV-19/mink/Netherlands/NB-EMC-55-19/2020\|EPI_ISL | ···················· | ···················· | ···················· | ···················· | ·····T·············· | ···················· | |
| Q | 8272 hCoV-19/Netherlands/LI-EMC-93/2020\|EPI_ISL_632431\| | ···················· | ···················· | ···················· | ···················· | ·····T·············· | ···················· | |
| Q | 8273 hCoV-19/Netherlands/LI-EMC-94/2020\|EPI_ISL_632432\| | ···················· | ···················· | ···················· | ···················· | ·····T·············· | ···················· | |
| Q | 8274 hCoV-19/Netherlands/LI-EMC-95/2020\|EPI_ISL_632433\| | ···················· | ···················· | ···················· | ···················· | ·····T·············· | ···················· | |
| Q | 8275 hCoV-19/mink/Netherlands/NB-EMC-62-2/2020\|EPI_ISL_ | ···················· | ···················· | ···················· | ···················· | ·····T·············· | ···················· | |
| Q | 8276 hCoV-19/mink/Netherlands/NB-EMC-62-3/2020\|EPI_ISL_ | ···················· | ···················· | ···················· | ···················· | ·····T·············· | ···················· | |
| Q | 8277 hCoV-19/mink/Netherlands/NB-EMC-62-4/2020\|EPI_ISL_ | ···················· | ···················· | ···················· | ···················· | ·····T·············· | ···················· | |
| Q | 8278 hCoV-19/mink/Netherlands/NB-EMC-62-5/2020\|EPI_ISL_ | ···················· | ···················· | ···················· | ···················· | ·····T·············· | ···················· | |
| Q | 8279 hCoV-19/mink/Netherlands/NB-EMC-62-6/2020\|EPI_ISL_ | ···················· | ···················· | ···················· | ···················· | ·····T·············· | ···················· | |
| Q | 8280 hCoV-19/mink/Netherlands/NB-EMC-64-5/2020\|EPI_ISL_ | ···················· | ···················· | ···················· | ···················· | ·····T·············· | ···················· | |
| Q | 8281 hCoV-19/Netherlands/GE-EMC-311/2020\|EPI_ISL_632440 | ···················· | ···················· | ···················· | ···················· | ···················· | ···················· | |
| Q | 8282 hCoV-19/Netherlands/ZH-EMC-795/2020\|EPI_ISL_632441 | ···················· | ···················· | ···················· | ···················· | ···················· | ···················· | |
| Q | 8283 hCoV-19/Netherlands/ZH-EMC-796/2020\|EPI_ISL_632442 | ···················· | ···················· | ···················· | ···················· | ···················· | ···················· | |
| Q | 8284 hCoV-19/Netherlands/ZH-EMC-810/2020\|EPI_ISL_632443 | ···················· | ···················· | ···················· | ···················· | ···················· | ···················· | |
| Q | 8285 hCoV-19/Netherlands/ZH-EMC-811/2020\|EPI_ISL_632444 | ···················· | ···················· | ···················· | ···················· | ···················· | ···················· | |
| Q | 8286 hCoV-19/Netherlands/FL-EMC-35/2020\|EPI_ISL_632445\| | ···················· | ···················· | ···················· | ···················· | ···················· | ···················· | |
| Q | 8287 hCoV-19/Netherlands/ZH-EMC-866/2020\|EPI_ISL_632448 | ···················· | ···················· | ···················· | ···················· | ···················· | ···················· | |
| Q | 8288 hCoV-19/mink/Netherlands/NB-EMC-54-1/2020\|EPI_ISL_ | ···················· | ···················· | ···················· | ···················· | ·····T·············· | ···················· | |
| Q | 8289 hCoV-19/mink/Netherlands/NB-EMC-54-2/2020\|EPI_ISL_ | ···················· | ···················· | ···················· | ···················· | ·····T·············· | ···················· | |
| Q | 8290 hCoV-19/mink/Netherlands/NB-EMC-54-5/2020\|EPI_ISL_ | ···················· | ···················· | ···················· | ···················· | ·····T·············· | ···················· | |
| Q | 8291 hCoV-19/mink/Netherlands/NB-EMC-56-6/2020\|EPI_ISL_ | ···················· | ···················· | ···················· | ···················· | ·····T·············· | ···················· | |
| Q | 8292 hCoV-19/mink/Netherlands/NB-EMC-56-7/2020\|EPI_ISL_ | ···················· | ···················· | ···················· | ···················· | ·····T·············· | ···················· | |
| Q | 8293 hCoV-19/mink/Netherlands/NB-EMC-56-10/2020\|EPI_ISL | ···················· | ···················· | ···················· | ···················· | ·····T·············· | ···················· | |
| Q | 8294 hCoV-19/mink/Netherlands/NB-EMC-57-1/2020\|EPI_ISL_ | ···················· | ···················· | ···················· | ···················· | ·····T·············· | ···················· | |
| Q | 8295 hCoV-19/mink/Netherlands/NB-EMC-57-2/2020\|EPI_ISL_ | ···················· | ···················· | ···················· | ···················· | ·····T·············· | ···················· | |
| Q | 8296 hCoV-19/England/QEUH-AD5D8B/2020\|EPI_ISL_633701\|20 | ···················· | ···················· | ···················· | ···················· | ···················· | ···················· | |

| | | | | | | | | |
|---|---|---|---|---|---|---|---|---|
| Q | 8297 | hCoV-19/England/QEUH-AD4EF2/2020\|EPI_ISL_633702\|20 | ···················· | ···················· | ···················· | ···················· | ···················· | ···················· |
| Q | 8298 | hCoV-19/England/QEUH-AD593C/2020\|EPI_ISL_633703\|20 | ···················· | ···················· | ···················· | ···················· | ···················· | ···················· |
| Q | 8299 | hCoV-19/England/QEUH-AD1244/2020\|EPI_ISL_633704\|20 | ···················· | ···················· | ···················· | ···················· | ···················· | ···················· |
| Q | 8300 | hCoV-19/England/QEUH-AD4E89/2020\|EPI_ISL_633705\|20 | ···················· | ···················· | ···················· | ···················· | ···················· | ···················· |
| Q | 8301 | hCoV-19/England/QEUH-AD0A6B/2020\|EPI_ISL_633707\|20 | ···················· | ···················· | ···················· | ···················· | ···················· | ···················· |
| Q | 8302 | hCoV-19/England/QEUH-AD0AC5/2020\|EPI_ISL_633709\|20 | ···················· | ···················· | ···················· | ···················· | ···················· | ···················· |
| Q | 8303 | hCoV-19/England/QEUH-AD5D7C/2020\|EPI_ISL_633710\|20 | ···················· | ···················· | ···················· | ···················· | ···················· | ···················· |
| Q | 8304 | hCoV-19/England/QEUH-AD5565/2020\|EPI_ISL_633711\|20 | ···················· | ·A·················· | ···················· | ···················· | ···················· | ···················· |
| Q | 8305 | hCoV-19/England/QEUH-AD1B85/2020\|EPI_ISL_633712\|20 | ···················· | ···················· | ···················· | ···················· | ···················· | ···················· |
| Q | 8306 | hCoV-19/England/QEUH-AD0AD4/2020\|EPI_ISL_633713\|20 | ···················· | ···················· | ···················· | ···················· | ···················· | ···················· |
| Q | 8307 | hCoV-19/England/QEUH-AD0A7A/2020\|EPI_ISL_633714\|20 | ···················· | ···················· | ···················· | ···················· | ···················· | ···················· |
| Q | 8308 | hCoV-19/England/QEUH-AD0A89/2020\|EPI_ISL_633715\|20 | ···················· | ···················· | ···················· | ···················· | ······T············· | ···················· |
| Q | 8309 | hCoV-19/England/QEUH-AD3655/2020\|EPI_ISL_633716\|20 | ···················· | ···················· | ···················· | ···················· | ···················· | ···················· |
| Q | 8310 | hCoV-19/England/QEUH-AD138D/2020\|EPI_ISL_633717\|20 | ···················· | ···················· | ···················· | ···················· | ···················· | ···················· |
| Q | 8311 | hCoV-19/England/QEUH-AD0B2C/2020\|EPI_ISL_633718\|20 | ···················· | ···················· | ···················· | ···················· | ···················· | ···················· |
| Q | 8312 | hCoV-19/England/QEUH-AD147B/2020\|EPI_ISL_633719\|20 | ···················· | ···················· | ···················· | ···················· | ···················· | ···················· |
| Q | 8313 | hCoV-19/England/QEUH-AD0DF9/2020\|EPI_ISL_633720\|20 | ···················· | ···················· | ···················· | ···················· | ···················· | ···················· |
| Q | 8314 | hCoV-19/England/QEUH-AD0A2F/2020\|EPI_ISL_633721\|20 | ···················· | ···················· | ···················· | ···················· | ···················· | ···················· |
| Q | 8315 | hCoV-19/England/QEUH-AD5987/2020\|EPI_ISL_633722\|20 | ···················· | ···················· | ···················· | ···················· | ···················· | ···················· |
| Q | 8316 | hCoV-19/England/QEUH-AD550B/2020\|EPI_ISL_633734\|20 | ···················· | ···················· | ···················· | ···················· | ···················· | ···················· |
| Q | 8317 | hCoV-19/England/QEUH-AD3190/2020\|EPI_ISL_633735\|20 | ···················· | ···················· | ···················· | ···················· | ···················· | ··A················· |
| Q | 8318 | hCoV-19/England/QEUH-AD4E10/2020\|EPI_ISL_633736\|20 | ···················· | ···················· | ···················· | ···················· | ···················· | ···················· |
| Q | 8319 | hCoV-19/England/QEUH-AD38C8/2020\|EPI_ISL_633737\|20 | ···················· | ···················· | ···················· | ···················· | ···················· | ···················· |
| Q | 8320 | hCoV-19/England/QEUH-AD0A98/2020\|EPI_ISL_633738\|20 | ···················· | ···················· | ···················· | ···················· | ···················· | ···················· |
| Q | 8321 | hCoV-19/England/QEUH-AD40A1/2020\|EPI_ISL_633739\|20 | ···················· | ···················· | ···················· | ···················· | ···················· | ···················· |
| Q | 8322 | hCoV-19/England/QEUH-AD55ED/2020\|EPI_ISL_633740\|20 | ···················· | ···················· | ···················· | ···················· | ···················· | ···················· |
| Q | 8323 | hCoV-19/England/QEUH-AD3163/2020\|EPI_ISL_633741\|20 | ···················· | ···················· | ···················· | ···················· | ···················· | ···················· |
| Q | 8324 | hCoV-19/England/QEUH-AD3251/2020\|EPI_ISL_633742\|20 | ···················· | ···················· | ···················· | ···················· | ···················· | ···················· |
| Q | 8325 | hCoV-19/England/QEUH-AD3260/2020\|EPI_ISL_633743\|20 | ···················· | ···················· | ···················· | ···················· | ···················· | ···················· |
| Q | 8326 | hCoV-19/England/QEUH-AD1B1C/2020\|EPI_ISL_633744\|20 | ···················· | ···················· | ···················· | ···················· | ···················· | ···················· |
| Q | 8327 | hCoV-19/England/QEUH-AD11A1/2020\|EPI_ISL_633745\|20 | ···················· | ···················· | ···················· | ···················· | ···················· | ···················· |
| Q | 8328 | hCoV-19/England/QEUH-AD385F/2020\|EPI_ISL_633746\|20 | ···················· | ···················· | ···················· | ···················· | ···················· | ···················· |
| Q | 8329 | hCoV-19/England/QEUH-AD31EB/2020\|EPI_ISL_633747\|20 | ···················· | ···················· | ···················· | ···················· | ···················· | ···················· |
| Q | 8330 | hCoV-19/England/QEUH-AD28D8/2020\|EPI_ISL_633748\|20 | ···················· | ·A·················· | ···················· | ···················· | ···················· | ···················· |
| Q | 8331 | hCoV-19/England/QEUH-AD5D6D/2020\|EPI_ISL_633749\|20 | ···················· | ···················· | ···················· | ···················· | ···················· | ···················· |
| Q | 8332 | hCoV-19/England/QEUH-AD1262/2020\|EPI_ISL_633750\|20 | ···················· | ···················· | ···················· | ···················· | ···················· | ···················· |
| Q | 8333 | hCoV-19/England/QEUH-AD5547/2020\|EPI_ISL_633752\|20 | ···················· | ···················· | ···················· | ···················· | ···················· | ···················· |
| Q | 8334 | hCoV-19/England/QEUH-AD0E6F/2020\|EPI_ISL_633753\|20 | ···················· | ···················· | ···················· | ···················· | ···················· | ···················· |
| Q | 8335 | hCoV-19/England/QEUH-AD4DE6/2020\|EPI_ISL_633754\|20 | ···················· | ···················· | ···················· | ···················· | ···················· | ···················· |
| Q | 8336 | hCoV-19/England/QEUH-AD351C/2020\|EPI_ISL_633755\|20 | ···················· | ···················· | ···················· | ···················· | ···················· | ···················· |
| Q | 8337 | hCoV-19/England/QEUH-AD59D2/2020\|EPI_ISL_633756\|20 | ···················· | ···················· | ···················· | ···················· | ···················· | ···················· |
| Q | 8338 | hCoV-19/England/QEUH-AD1B94/2020\|EPI_ISL_633757\|20 | ···················· | ···················· | ···················· | ···················· | ···················· | ···················· |
| Q | 8339 | hCoV-19/England/QEUH-AD32D9/2020\|EPI_ISL_633758\|20 | ···················· | ···················· | ···················· | ···················· | ···················· | ···················· |
| Q | 8340 | hCoV-19/England/QEUH-AD59F0/2020\|EPI_ISL_633761\|20 | ···················· | ···················· | ···················· | ···················· | ···················· | ···················· |
| Q | 8341 | hCoV-19/England/QEUH-AD337C/2020\|EPI_ISL_633762\|20 | ···················· | ·A·················· | ···················· | ···················· | ···················· | ··A················· |
| Q | 8342 | hCoV-19/England/QEUH-AD3628/2020\|EPI_ISL_633763\|20 | ···················· | ···················· | ···················· | ···················· | ···················· | ···················· |
| Q | 8343 | hCoV-19/England/QEUH-AD0DBD/2020\|EPI_ISL_633764\|20 | ···················· | ···················· | ···················· | ···················· | ···················· | ···················· |
| Q | 8344 | hCoV-19/England/QEUH-AD4EB6/2020\|EPI_ISL_633765\|20 | ···················· | ···················· | ···················· | ···················· | ···················· | ···················· |
| Q | 8345 | hCoV-19/England/QEUH-AD160C/2020\|EPI_ISL_633766\|20 | ···················· | ···················· | ···················· | ···················· | ······T············· | ···················· |
| Q | 8346 | hCoV-19/England/QEUH-AD0825/2020\|EPI_ISL_633767\|20 | ···················· | ···················· | ···················· | ···················· | ···················· | ···················· |
| Q | 8347 | hCoV-19/England/QEUH-AD3181/2020\|EPI_ISL_633768\|20 | ···················· | ···················· | ···················· | ···················· | ···················· | ···················· |
| Q | 8348 | hCoV-19/England/QEUH-AD1217/2020\|EPI_ISL_633769\|20 | ···················· | ···················· | ···················· | ···················· | ···················· | ···················· |
| Q | 8349 | hCoV-19/England/QEUH-AD1411/2020\|EPI_ISL_633770\|20 | ···················· | ···················· | ···················· | ···················· | ···················· | ···················· |
| Q | 8350 | hCoV-19/England/QEUH-AD1B49/2020\|EPI_ISL_633771\|20 | ···················· | ···················· | ···················· | ···················· | ···················· | ···················· |
| Q | 8351 | hCoV-19/England/QEUH-AD15C3/2020\|EPI_ISL_633772\|20 | ···················· | ···················· | ···················· | ···················· | ···················· | ···················· |
| Q | 8352 | hCoV-19/England/QEUH-AD08DA/2020\|EPI_ISL_633773\|20 | ···················· | ···················· | ···················· | ···················· | ···················· | ···················· |
| Q | 8353 | hCoV-19/England/QEUH-AD162A/2020\|EPI_ISL_633774\|20 | ···················· | ···················· | ···················· | ···················· | ···················· | ···················· |
| Q | 8354 | hCoV-19/England/QEUH-AD3567/2020\|EPI_ISL_633775\|20 | ···················· | ···················· | ···················· | ···················· | ···················· | ···················· |
| Q | 8355 | hCoV-19/England/QEUH-AD38D7/2020\|EPI_ISL_633776\|20 | ···················· | ···················· | ···················· | ···················· | ···················· | ···················· |
| Q | 8356 | hCoV-19/England/QEUH-AD353A/2020\|EPI_ISL_633778\|20 | ···················· | ···················· | ···················· | ···················· | ···················· | ···················· |
| Q | 8357 | hCoV-19/England/QEUH-AD0AF2/2020\|EPI_ISL_633779\|20 | ···················· | ···················· | ···················· | ···················· | ···················· | ···················· |
| Q | 8358 | hCoV-19/England/QEUH-AD3637/2020\|EPI_ISL_633780\|20 | ···················· | ···················· | ···················· | ···················· | ···················· | ···················· |
| Q | 8359 | hCoV-19/England/QEUH-AD5556/2020\|EPI_ISL_633783\|20 | ···················· | ·A·················· | ···················· | ···················· | ···················· | ···················· |
| Q | 8360 | hCoV-19/England/QEUH-AD35EF/2020\|EPI_ISL_633784\|20 | ···················· | ···················· | ···················· | ···················· | ···················· | ···················· |
| Q | 8361 | hCoV-19/England/QEUH-AD0A4D/2020\|EPI_ISL_633785\|20 | ···················· | ···················· | ···················· | ···················· | ···················· | ···················· |
| Q | 8362 | hCoV-19/England/QEUH-AD07CE/2020\|EPI_ISL_633787\|20 | ···················· | ···················· | ···················· | ···················· | ···················· | ···················· |
| Q | 8363 | hCoV-19/England/QEUH-AD10FF/2020\|EPI_ISL_633801\|20 | ···················· | ···················· | ···················· | ···················· | ···················· | ···················· |
| Q | 8364 | hCoV-19/England/QEUH-AD4F77/2020\|EPI_ISL_633802\|20 | ···················· | ···················· | ···················· | ···················· | ···················· | ···················· |
| Q | 8365 | hCoV-19/England/QEUH-AD38E6/2020\|EPI_ISL_633803\|20 | ···················· | ···················· | ···················· | ···················· | ···················· | ···················· |
| Q | 8366 | hCoV-19/England/QEUH-AD5DB8/2020\|EPI_ISL_633804\|20 | ···················· | ···················· | ···················· | ···················· | ···················· | ···················· |
| Q | 8367 | hCoV-19/England/QEUH-AD32BB/2020\|EPI_ISL_633805\|20 | ···················· | ···················· | ···················· | ···················· | ···················· | ···················· |
| Q | 8368 | hCoV-19/England/QEUH-AD3224/2020\|EPI_ISL_633807\|20 | ···················· | ···················· | ···················· | ···················· | ···················· | ···················· |
| Q | 8369 | hCoV-19/England/QEUH-AD3989/2020\|EPI_ISL_633808\|20 | ···················· | ···················· | ···················· | ···················· | ···················· | ···················· |
| Q | 8370 | hCoV-19/England/QEUH-AD1226/2020\|EPI_ISL_633810\|20 | ···················· | ···················· | ···················· | ···················· | ···················· | ···················· |
| Q | 8371 | hCoV-19/England/QEUH-AD11FC/2020\|EPI_ISL_633811\|20 | ···················· | ···················· | ···················· | ···················· | ···················· | ···················· |
| Q | 8372 | hCoV-19/England/QEUH-AD3127/2020\|EPI_ISL_633812\|20 | ···················· | ···················· | ···················· | ···················· | ···················· | ···················· |
| Q | 8373 | hCoV-19/England/QEUH-AD1569/2020\|EPI_ISL_633816\|20 | ···················· | ···················· | ···················· | ···················· | ···················· | ···················· |
| Q | 8374 | hCoV-19/England/QEUH-AD152D/2020\|EPI_ISL_633817\|20 | ···················· | ·A·················· | ···················· | ···················· | ···················· | ··A················· |
| Q | 8375 | hCoV-19/England/QEUH-AD28BA/2020\|EPI_ISL_633818\|20 | ···················· | ···················· | ···················· | ···················· | ···················· | ···················· |
| Q | 8376 | hCoV-19/England/QEUH-AD31CD/2020\|EPI_ISL_633819\|20 | ···················· | ···················· | ···················· | ···················· | ···················· | ···················· |

Q 8377 hCoV-19/England/QEUH-AD0BB3/2020|EPI_ISL_633820|20 ···················· ···················· ···················· ···················· ···················· ····················
Q 8378 hCoV-19/USA/CA-ALSR-3428/2020|EPI_ISL_636022|2020- ···················· ·A·················· ···················· ···················· ···················· ····················
Q 8379 hCoV-19/USA/CA-ALSR-3429/2020|EPI_ISL_636023|2020- ···················· ···················· ···················· ···················· ···················· ····················
Q 8380 hCoV-19/USA/CA-ALSR-3430/2020|EPI_ISL_636024|2020- ···················· ·A·················· ···················· ···················· ···················· ····················
Q 8381 hCoV-19/USA/CA-ALSR-3432/2020|EPI_ISL_636025|2020- ···················· ···················· ···················· ···················· ···················· ····················
Q 8382 hCoV-19/USA/CA-ALSR-3434/2020|EPI_ISL_636026|2020- ···················· ·A·················· ···················· ···················· ···················· ····················
Q 8383 hCoV-19/USA/CA-ALSR-3435/2020|EPI_ISL_636027|2020- ···················· ·A·················· ···················· ···················· ···················· ····················
Q 8384 hCoV-19/USA/CA-ALSR-3436/2020|EPI_ISL_636028|2020- ···················· ·A·················· ···················· ···················· ···················· ····················
Q 8385 hCoV-19/USA/CA-ALSR-3437/2020|EPI_ISL_636029|2020- ···················· ·A·················· ···················· ···················· ···················· ····················
Q 8386 hCoV-19/USA/CA-ALSR-3441/2020|EPI_ISL_636031|2020- ···················· ·A·················· ···················· ···················· ···················· ····················
Q 8387 hCoV-19/USA/CA-ALSR-3442/2020|EPI_ISL_636032|2020- ···················· ·A·················· ···················· ···················· ···················· ····················
Q 8388 hCoV-19/USA/CA-ALSR-3443/2020|EPI_ISL_636033|2020- ···················· ···················· ···················· ···················· ···················· ····················
Q 8389 hCoV-19/USA/CA-ALSR-3445/2020|EPI_ISL_636034|2020- ···················· ···················· ···················· ···················· ···················· ····················
Q 8390 hCoV-19/USA/CA-ALSR-3448/2020|EPI_ISL_636035|2020- ···················· ···················· ···················· ···················· ···················· ····················
Q 8391 hCoV-19/USA/CA-ALSR-3451/2020|EPI_ISL_636036|2020- ···················· ·A·················· ···················· ···················· ···················· ····················
Q 8392 hCoV-19/USA/CA-ALSR-3453/2020|EPI_ISL_636037|2020- ···················· ···················· ···················· ···················· ···················· ····················
Q 8393 hCoV-19/USA/CA-ALSR-3458/2020|EPI_ISL_636038|2020- ···················· ·A·················· ···················· ···················· ···················· ····················
Q 8394 hCoV-19/USA/CA-ALSR-3460/2020|EPI_ISL_636039|2020- ···················· ·A·················· ···················· ···················· ···················· ····················
Q 8395 hCoV-19/USA/CA-ALSR-3463/2020|EPI_ISL_636040|2020- ···················· ·A·················· ···················· ···················· ···················· ····················
Q 8396 hCoV-19/USA/CA-ALSR-3464/2020|EPI_ISL_636041|2020- ···················· ···················· ···················· ···················· ···················· ····················
Q 8397 hCoV-19/USA/CA-ALSR-3470/2020|EPI_ISL_636042|2020- ···················· ·A·················· ···················· ···················· ···················· ····················
Q 8398 hCoV-19/USA/CA-ALSR-3471/2020|EPI_ISL_636043|2020- ···················· ·A·················· ···················· ···················· ···A················ ····················
Q 8399 hCoV-19/USA/CA-ALSR-3472/2020|EPI_ISL_636044|2020- ···················· ·A·················· ···················· ···················· ···················· ····················
Q 8400 hCoV-19/USA/CA-ALSR-3473/2020|EPI_ISL_636045|2020- ···················· ·A·················· ···················· ···················· ···················· ····················
Q 8401 hCoV-19/USA/CA-ALSR-3474/2020|EPI_ISL_636046|2020- ···················· ···················· ···················· ···················· ···················· ····················
Q 8402 hCoV-19/USA/CA-ALSR-3478/2020|EPI_ISL_636047|2020- ···················· ·A·················· ···················· ···················· ···················· ····················
Q 8403 hCoV-19/USA/CA-ALSR-3479/2020|EPI_ISL_636048|2020- ···················· ·A·················· ···················· ···················· ···················· ····················
Q 8404 hCoV-19/USA/CA-ALSR-3480/2020|EPI_ISL_636049|2020- ···················· ···················· ···················· ···················· ···················· ····················
Q 8405 hCoV-19/USA/CA-ALSR-3494/2020|EPI_ISL_636050|2020- ···················· ·A·················· ···················· ···················· ···················· ····················
Q 8406 hCoV-19/USA/CA-ALSR-3495/2020|EPI_ISL_636051|2020- ···················· ·A·················· ···················· ···················· ···················· ····················
Q 8407 hCoV-19/USA/CA-ALSR-3496/2020|EPI_ISL_636052|2020- ···················· ·A·················· ···················· ···················· ···················· ····················
Q 8408 hCoV-19/USA/CA-ALSR-3497/2020|EPI_ISL_636053|2020- ···················· ·A·················· ···················· ···················· ···················· ····················
Q 8409 hCoV-19/USA/CA-ALSR-3498/2020|EPI_ISL_636054|2020- ···················· ·A·················· ···················· ···················· ···················· ····················
Q 8410 hCoV-19/USA/CA-ALSR-3501/2020|EPI_ISL_636056|2020- ···················· ···················· ···················· ···················· ···················· ····················
Q 8411 hCoV-19/USA/CA-ALSR-3505/2020|EPI_ISL_636057|2020- ···················· ·A·················· ···················· ···················· ···················· ····················
Q 8412 hCoV-19/USA/CA-ALSR-3507/2020|EPI_ISL_636059|2020- ···················· ·A·················· ···················· ···················· ···················· ····················
Q 8413 hCoV-19/USA/CA-ALSR-3510/2020|EPI_ISL_636060|2020- ···················· ···················· ···················· ···················· ···················· ····················
Q 8414 hCoV-19/USA/CA-ALSR-3512/2020|EPI_ISL_636061|2020- ···················· ···················· ···················· ···················· ···················· ····················
Q 8415 hCoV-19/USA/CA-ALSR-3514/2020|EPI_ISL_636062|2020- ·······T············ ·A·················· ···················· ···················· ···················· ····················
Q 8416 hCoV-19/USA/CA-ALSR-3516/2020|EPI_ISL_636063|2020- ···················· ·A·················· ···················· ···················· ···················· ····················
Q 8417 hCoV-19/USA/CA-ALSR-3519/2020|EPI_ISL_636065|2020- ···················· ·A·················· ···················· ···················· ···················· ····················
Q 8418 hCoV-19/USA/CA-ALSR-3521/2020|EPI_ISL_636066|2020- ···················· ·A·················· ···················· ···················· ···················· ····················
Q 8419 hCoV-19/USA/CA-ALSR-3522/2020|EPI_ISL_636067|2020- ···················· ·A·················· ···················· ···················· ···················· ····················
Q 8420 hCoV-19/USA/CA-ALSR-3524/2020|EPI_ISL_636068|2020- ·······T············ ·A·················· ···················· ···················· ···················· ····················
Q 8421 hCoV-19/USA/CA-ALSR-3526/2020|EPI_ISL_636069|2020- ···················· ·A·················· ···················· ···················· ···················· ····················
Q 8422 hCoV-19/USA/CA-ALSR-3527/2020|EPI_ISL_636070|2020- ···················· ·A·················· ···················· ···················· ···················· ····················
Q 8423 hCoV-19/USA/CA-ALSR-3528/2020|EPI_ISL_636071|2020- ···················· ·A·················· ···················· ···················· ···················· ····················
Q 8424 hCoV-19/USA/CA-ALSR-3532/2020|EPI_ISL_636072|2020- ···················· ·A·················· ···················· ···················· ···················· ····················
Q 8425 hCoV-19/USA/CA-ALSR-3533/2020|EPI_ISL_636073|2020- ···················· ·A·················· ···················· ···················· ···················· ····················
Q 8426 hCoV-19/USA/CA-ALSR-3535/2020|EPI_ISL_636074|2020- ···················· ·A·················· ···················· ···················· ···················· ····················
Q 8427 hCoV-19/USA/CA-ALSR-3539/2020|EPI_ISL_636075|2020- ···················· ···················· ···················· ···················· ···················· ····················
Q 8428 hCoV-19/USA/CA-ALSR-3541/2020|EPI_ISL_636076|2020- ···················· ···················· ···················· ···················· ···················· ····················
Q 8429 hCoV-19/USA/CA-ALSR-3544/2020|EPI_ISL_636077|2020- ···················· ·A·················· ···················· ···················· ···················· ····················
Q 8430 hCoV-19/USA/CA-ALSR-3545/2020|EPI_ISL_636078|2020- ···················· ·A·················· ···················· ···················· ···················· ····················
Q 8431 hCoV-19/USA/CA-ALSR-3551/2020|EPI_ISL_636079|2020- ···················· ·A·················· ···················· ···················· ···················· ····················
Q 8432 hCoV-19/USA/CA-ALSR-3554/2020|EPI_ISL_636080|2020- ···················· ·A·················· ···················· ···················· ···················· ····················
Q 8433 hCoV-19/USA/CA-ALSR-3555/2020|EPI_ISL_636081|2020- ···················· ·A·················· ···················· ···················· ···················· ····················
Q 8434 hCoV-19/USA/CA-ALSR-3557/2020|EPI_ISL_636082|2020- ···················· ·A·················· ···················· ···················· ···················· ····················
Q 8435 hCoV-19/USA/CA-ALSR-3560/2020|EPI_ISL_636084|2020- ···················· ·A·················· ···················· ···················· ···················· ····················
Q 8436 hCoV-19/USA/CA-ALSR-3563/2020|EPI_ISL_636085|2020- ···················· ·A·················· ···················· ···················· ···················· ····················
Q 8437 hCoV-19/USA/CA-ALSR-3566/2020|EPI_ISL_636086|2020- ···················· ·A·················· ···················· ···················· ···················· ····················
Q 8438 hCoV-19/USA/CA-ALSR-3567/2020|EPI_ISL_636087|2020- ···················· ·A·················· ···················· ···················· ···················· ····················
Q 8439 hCoV-19/USA/CA-ALSR-3569/2020|EPI_ISL_636088|2020- ···················· ·A·················· ···················· ···················· ···················· ····················
Q 8440 hCoV-19/USA/CA-ALSR-3572/2020|EPI_ISL_636090|2020- ···················· ·A·················· ···················· ···················· ···················· ····················
Q 8441 hCoV-19/USA/CA-ALSR-3578/2020|EPI_ISL_636091|2020- ···················· ·A·················· ···················· ···················· ···················· ····················
Q 8442 hCoV-19/USA/CA-ALSR-3579/2020|EPI_ISL_636092|2020- ···················· ·A·················· ···················· ···················· ···················· ····················
Q 8443 hCoV-19/USA/CA-ALSR-3580/2020|EPI_ISL_636093|2020- ···················· ···················· ···················· ···················· ···················· ····················
Q 8444 hCoV-19/USA/CA-ALSR-3581/2020|EPI_ISL_636094|2020- ···················· ·A·················· ···················· ···················· ···················· ····················
Q 8445 hCoV-19/USA/CA-ALSR-3582/2020|EPI_ISL_636095|2020- ···················· ·A·················· ···················· ···················· ···················· ····················
Q 8446 hCoV-19/USA/CA-ALSR-3583/2020|EPI_ISL_636096|2020- ···················· ·A·················· ···················· ···················· ···················· ····················
Q 8447 hCoV-19/USA/CA-ALSR-3585/2020|EPI_ISL_636097|2020- ···················· ·A·················· ···················· ···················· ···················· ····················
Q 8448 hCoV-19/USA/CA-ALSR-3589/2020|EPI_ISL_636098|2020- ···················· ···················· ···················· ···················· ···················· ····················
Q 8449 hCoV-19/Czech Republic/NRL_9678/2020|EPI_ISL_62660 ···················· ···················· ···················· ···················· ···················· ····················
Q 8450 hCoV-19/Czech Republic/NRL_9679/2020|EPI_ISL_62660 ···················· ···················· ···················· ···················· ···················· ····················
Q 8451 hCoV-19/Czech Republic/NRL_9681/2020|EPI_ISL_62660 ···················· ···················· ···················· ···················· ···················· ····················
Q 8452 hCoV-19/Czech Republic/NRL_9682/2020|EPI_ISL_62660 ···················· ···················· ···················· ···················· ···················· ····················
Q 8453 hCoV-19/Czech Republic/NRL_9703/2020|EPI_ISL_62660 ···················· ···················· ···················· ···················· ···················· ····················
Q 8454 hCoV-19/Czech Republic/NRL_9713/2020|EPI_ISL_62660 ···················· ···················· ···················· ···················· ···················· ····················
Q 8455 hCoV-19/Czech Republic/NRL_9743/2020|EPI_ISL_62660 ···················· ···················· ···················· ···················· ···················· ····················
Q 8456 hCoV-19/Czech Republic/NRL_9818/2020|EPI_ISL_62660 ···················· ···················· ···················· ···················· ···················· ····················

Q 8457 hCoV-19/Czech Republic/NRL_9837/2020|EPI_ISL_62660 ···················· ···················· ····················· ···················· ···················· ····················
Q 8458 hCoV-19/Czech Republic/NRL_9840/2020|EPI_ISL_62660 ···················· ···················· ····················· ···················· ···················· ····················
Q 8459 hCoV-19/Czech Republic/NRL_9875/2020|EPI_ISL_62661 ···················· ···················· ····················· ···················· ···················· ····················
Q 8460 hCoV-19/Czech Republic/NRL_9880/2020|EPI_ISL_62661 ···················· ···················· ····················· ···················· ···················· ····················
Q 8461 hCoV-19/Czech Republic/NRL_9881/2020|EPI_ISL_62661 ···················· ···················· ····················· ···················· ···················· ····················
Q 8462 hCoV-19/Czech Republic/NRL_9882/2020|EPI_ISL_62661 ···················· ···················· ····················· ···················· ···················· ····················
Q 8463 hCoV-19/Czech Republic/NRL_9883/2020|EPI_ISL_62661 ···················· ···················· ····················· ···················· ···················· ····················
Q 8464 hCoV-19/Czech Republic/NRL_9885/2020|EPI_ISL_62661 ···················· ···················· ····················· ···················· ···················· ····················
Q 8465 hCoV-19/Czech Republic/NRL_9887/2020|EPI_ISL_62661 ···················· ···················· ····················· ···················· ···················· ····················
Q 8466 hCoV-19/Wales/PHWC-483B79/2020|EPI_ISL_627900|2020 ···················· ···················· ····················· ···················· ···················· ····················
Q 8467 hCoV-19/Wales/PHWC-483B88/2020|EPI_ISL_627901|2020 ···················· ···················· ····················· ···················· ···················· ····················
Q 8468 hCoV-19/Wales/PHWC-483B97/2020|EPI_ISL_627902|2020 ···················· ···················· ····················· ···················· ···················· ····················
Q 8469 hCoV-19/Wales/PHWC-483BA6/2020|EPI_ISL_627903|2020 ···················· ···················· ····················· ···················· ···················· ····················
Q 8470 hCoV-19/Wales/PHWC-483BB5/2020|EPI_ISL_627904|2020 ···················· ···················· ····················· ···················· ···················· ····················
Q 8471 hCoV-19/Wales/PHWC-483BC4/2020|EPI_ISL_627905|2020 ···················· ···················· ····················· ···················· ···················· ····················
Q 8472 hCoV-19/Wales/PHWC-483BD3/2020|EPI_ISL_627906|2020 ···················· ···················· ····················· ···················· ···················· ····················
Q 8473 hCoV-19/Wales/PHWC-483BE2/2020|EPI_ISL_627907|2020 ···················· ···················· ····················· ···················· ···················· ····················
Q 8474 hCoV-19/Wales/PHWC-483BF1/2020|EPI_ISL_627908|2020 ···················· ···················· ····················· ···················· ···················· ····················
Q 8475 hCoV-19/Wales/PHWC-483C0D/2020|EPI_ISL_627909|2020 ···················· ···················· ····················· ···················· ···················· ····················
Q 8476 hCoV-19/Wales/PHWC-483C1C/2020|EPI_ISL_627910|2020 ···················· ···················· ····················· ···················· ···················· ····················
Q 8477 hCoV-19/Wales/PHWC-483C2B/2020|EPI_ISL_627911|2020 ···················· ···················· ····················· ···················· ···················· ····················
Q 8478 hCoV-19/Wales/PHWC-483C3A/2020|EPI_ISL_627912|2020 ···················· ···················· ····················· ···················· ···················· ····················
Q 8479 hCoV-19/Wales/PHWC-483C49/2020|EPI_ISL_627913|2020 ···················· ···················· ····················· ···················· ···················· ····················
Q 8480 hCoV-19/Wales/PHWC-483C58/2020|EPI_ISL_627914|2020 ···················· ···················· ····················· ···················· ···················· ····················
Q 8481 hCoV-19/Wales/PHWC-483C76/2020|EPI_ISL_627915|2020 ···················· ···················· ····················· ···················· ···················· ····················
Q 8482 hCoV-19/Wales/PHWC-483C85/2020|EPI_ISL_627916|2020 ···················· ···················· ····················· ···················· ···················· ····················
Q 8483 hCoV-19/Wales/PHWC-483C94/2020|EPI_ISL_627917|2020 ···················· ···················· ····················· ···················· ···················· ····················
Q 8484 hCoV-19/Wales/PHWC-483CA3/2020|EPI_ISL_627918|2020 ···················· ···················· ····················· ···················· ···················· ····················
Q 8485 hCoV-19/Wales/PHWC-483CB2/2020|EPI_ISL_627919|2020 ···················· ···················· ····················· ···················· ···················· ····················
Q 8486 hCoV-19/Wales/PHWC-483CD0/2020|EPI_ISL_627920|2020 ···················· ···················· ····················· ···················· ···················· ····················
Q 8487 hCoV-19/Wales/PHWC-483D0A/2020|EPI_ISL_627921|2020 ···················· ···················· ····················· ···················· ···················· ····················
Q 8488 hCoV-19/Wales/PHWC-483D19/2020|EPI_ISL_627922|2020 ···················· ···················· ····················· ···················· ···················· ····················
Q 8489 hCoV-19/Wales/PHWC-483D28/2020|EPI_ISL_627923|2020 ···················· ···················· ····················· ···················· ···················· ····················
Q 8490 hCoV-19/Wales/PHWC-483D37/2020|EPI_ISL_627924|2020 ···················· ···················· ····················· ···················· ···················· ····················
Q 8491 hCoV-19/Wales/PHWC-483D46/2020|EPI_ISL_627925|2020 ···················· ···················· ····················· ···················· ···················· ····················
Q 8492 hCoV-19/Wales/PHWC-483D64/2020|EPI_ISL_627926|2020 ···················· ···················· ····················· ···················· ···················· ····················
Q 8493 hCoV-19/Wales/PHWC-483D73/2020|EPI_ISL_627927|2020 ···················· ···················· ····················· ···················· ···A················ ····················
Q 8494 hCoV-19/Wales/PHWC-483D91/2020|EPI_ISL_627928|2020 ···················· ···················· ····················· ···················· ···················· ····················
Q 8495 hCoV-19/Wales/PHWC-483DA0/2020|EPI_ISL_627929|2020 ···················· ···················· ····················· ···················· ···················· ····················
Q 8496 hCoV-19/Wales/PHWC-483DCE/2020|EPI_ISL_627930|2020 ···················· ···················· ····················· ···················· ···················· ····················
Q 8497 hCoV-19/Wales/PHWC-483DEC/2020|EPI_ISL_627931|2020 ···················· ···················· ····················· ···················· ···················· ····················
Q 8498 hCoV-19/Wales/PHWC-483DFB/2020|EPI_ISL_627932|2020 ···················· ···················· ····················· ···················· ···················· ····················
Q 8499 hCoV-19/Wales/PHWC-483E07/2020|EPI_ISL_627933|2020 ···················· ···················· ····················· ···················· ···················· ····················
Q 8500 hCoV-19/Wales/PHWC-483E16/2020|EPI_ISL_627934|2020 ···················· ···················· ····················· ···················· ···················· ····················
Q 8501 hCoV-19/Wales/PHWC-483E25/2020|EPI_ISL_627935|2020 ···················· ···················· ····················· ···················· ···················· ····················
Q 8503 hCoV-19/Wales/PHWC-483E52/2020|EPI_ISL_627937|2020 ···················· ···················· ····················· ···················· ···················· ····················
Q 8505 hCoV-19/Wales/PHWC-483E70/2020|EPI_ISL_627939|2020 ···················· ···················· ····················· ···················· ···················· ····················
Q 8506 hCoV-19/Wales/PHWC-483E8F/2020|EPI_ISL_627940|2020 ···················· ···················· ····················· ···················· ···················· ····················
Q 8507 hCoV-19/Wales/PHWC-483E9E/2020|EPI_ISL_627941|2020 ···················· ···················· ····················· ···················· ···················· ····················
Q 8508 hCoV-19/Wales/PHWC-483EAD/2020|EPI_ISL_627942|2020 ···················· ···················· ····················· ···················· ···················· ····················
Q 8509 hCoV-19/Wales/PHWC-483EDA/2020|EPI_ISL_627943|2020 ···················· ···················· ····················· ···················· ···················· ····················
Q 8511 hCoV-19/Wales/PHWC-483F04/2020|EPI_ISL_627945|2020 ···················· ···················· ····················· ···················· ···················· ····················
Q 8512 hCoV-19/Wales/PHWC-483F22/2020|EPI_ISL_627946|2020 ···················· ···················· ····················· ···················· ···················· ····················
Q 8513 hCoV-19/Wales/PHWC-483F31/2020|EPI_ISL_627947|2020 ···················· ···················· ····················· ···················· ···················· ····················
Q 8514 hCoV-19/England/ALDP-ABD2E4/2020|EPI_ISL_631127|20 ···················· ···················· ····················· ···················· ···················· ····················
Q 8515 hCoV-19/England/ALDP-ABD25D/2020|EPI_ISL_631128|20 ···················· ···················· ····················· ···················· ···················· ····················
Q 8516 hCoV-19/England/CAMC-AAF50A/2020|EPI_ISL_631129|20 ···················· ···················· ····················· ···················· ···················· ····················
Q 8517 hCoV-19/England/MILK-ABBF76/2020|EPI_ISL_631130|20 ···················· ···················· ····················· ···················· ···················· ····················
Q 8518 hCoV-19/England/ALDP-AC5DD9/2020|EPI_ISL_631131|20 ···················· ···················· ····················· ···················· ···················· ····················
Q 8519 hCoV-19/England/ALDP-AC5E6D/2020|EPI_ISL_631133|20 ···················· ···················· ····················· ···················· ···················· ····················
Q 8520 hCoV-19/England/MILK-ABB84E/2020|EPI_ISL_631134|20 ···················· ···················· ····················· ···················· ······T············· ····················
Q 8521 hCoV-19/England/CAMC-AAF9FF/2020|EPI_ISL_631135|20 ···················· ···················· ····················· ···················· ···················· ····················
Q 8522 hCoV-19/England/MILK-ABBC06/2020|EPI_ISL_631136|20 ···················· ···················· ····················· ···················· ···················· ····················
Q 8523 hCoV-19/England/CAMC-AAF79B/2020|EPI_ISL_631137|20 ···················· ···················· ····················· ···················· ···················· ····················
Q 8524 hCoV-19/England/MILK-ABBBCD/2020|EPI_ISL_631138|20 ···················· ···················· ····················· ···················· ···················· ····················
Q 8525 hCoV-19/England/MILK-ABC133/2020|EPI_ISL_631139|20 ···················· ···················· ····················· ···················· ···················· ····················
Q 8526 hCoV-19/England/MILK-ABC115/2020|EPI_ISL_631140|20 ···················· ···················· ····················· ···················· ···················· ····················
Q 8527 hCoV-19/England/MILK-ABC1AC/2020|EPI_ISL_631141|20 ···················· ···················· ····················· ···················· ···················· ····················
Q 8528 hCoV-19/England/CAMC-AAFA47/2020|EPI_ISL_631142|20 ···················· ···················· ····················· ···················· ···················· ····················
Q 8529 hCoV-19/England/MILK-ABC40D/2020|EPI_ISL_631143|20 ···················· ···················· ····················· ···················· ···················· ····················
Q 8530 hCoV-19/England/CAMC-AAFA1A/2020|EPI_ISL_631144|20 ···················· ···················· ····················· ···················· ···················· ····················
Q 8531 hCoV-19/England/MILK-ABB8B7/2020|EPI_ISL_631145|20 ···················· ···················· ····················· ···················· ···················· ····················
Q 8532 hCoV-19/England/MILK-ABBD30/2020|EPI_ISL_631146|20 ···················· ···················· ····················· ···················· ···················· ····················
Q 8533 hCoV-19/England/CAMC-AAF616/2020|EPI_ISL_631148|20 ···················· ···················· ····················· ···················· ···················· ····················
Q 8534 hCoV-19/England/CAMC-AAFF66/2020|EPI_ISL_631149|20 ···················· ···················· ····················· ···················· ···················· ····················
Q 8535 hCoV-19/England/ALDP-AC5DAC/2020|EPI_ISL_631150|20 ···················· ···················· ····················· ···················· ···················· ····················
Q 8536 hCoV-19/England/MILK-ABC17F/2020|EPI_ISL_631151|20 ···················· ···················· ····················· ···················· ······T············· ····················
Q 8537 hCoV-19/England/MILK-ABC0BE/2020|EPI_ISL_631152|20 ···················· ···················· ····················· ···················· ···················· ····················
Q 8538 hCoV-19/England/MILK-ABBD21/2020|EPI_ISL_631153|20 ···················· ·A·················· ····················· ···················· ···················· ···A················
Q 8539 hCoV-19/England/CAMC-AAF9D1/2020|EPI_ISL_631154|20 ···················· ···················· ····················· ···················· ···················· ····················

| | | | | | | | |
|---|---|---|---|---|---|---|---|
| Q | 8540 hCoV-19/England/CAMC-AAF7AA/2020\|EPI_ISL_631155\|20 | ···················· | ···················· | ····················· | ···················· | ···················· | ···················· |
| Q | 8541 hCoV-19/England/MILK-ABBB90/2020\|EPI_ISL_631156\|20 | ···················· | ···················· | ····················· | ···················· | ···················· | ···················· |
| Q | 8542 hCoV-19/England/CAMC-AAFF57/2020\|EPI_ISL_631160\|20 | ···················· | ···················· | ····················· | ···················· | ···················· | ···················· |
| Q | 8543 hCoV-19/England/MILK-ABB86C/2020\|EPI_ISL_631161\|20 | ···················· | ···················· | ····················· | ···················· | ···················· | ···················· |
| Q | 8544 hCoV-19/England/MILK-ABB820/2020\|EPI_ISL_631162\|20 | ···················· | ···················· | ····················· | ···················· | ···················· | ···················· |
| Q | 8545 hCoV-19/England/ALDP-ABD24E/2020\|EPI_ISL_631163\|20 | ···················· | ···················· | ····················· | ···················· | ···················· | ···················· |
| Q | 8546 hCoV-19/England/CAMC-AAFF39/2020\|EPI_ISL_631164\|20 | ···················· | ···················· | ····················· | ···················· | ···················· | ···················· |
| Q | 8547 hCoV-19/England/ALDP-AC1292/2020\|EPI_ISL_631165\|20 | ···················· | ·A·················· | ····················· | ···················· | ···················· | ···················· |
| Q | 8548 hCoV-19/England/MILK-ABBDE5/2020\|EPI_ISL_631166\|20 | ···················· | ···················· | ····················· | ···················· | ···················· | ···················· |
| Q | 8549 hCoV-19/England/MILK-ABC4C1/2020\|EPI_ISL_631167\|20 | ···················· | ···················· | ····················· | ···················· | ·····T·············· | ···················· |
| Q | 8550 hCoV-19/England/MILK-ABC2A9/2020\|EPI_ISL_631168\|20 | ···················· | ·A·················· | ····················· | ···················· | ···················· | ··A················· |
| Q | 8551 hCoV-19/England/MILK-ABBEF1/2020\|EPI_ISL_631169\|20 | ···················· | ···················· | ····················· | ···················· | ···················· | ···················· |
| Q | 8552 hCoV-19/England/MILK-ABBF2B/2020\|EPI_ISL_631170\|20 | ···················· | ·A·················· | ····················· | ···················· | ···················· | ··A················· |
| Q | 8553 hCoV-19/England/CAMC-AAF91D/2020\|EPI_ISL_631171\|20 | ···················· | ···················· | ····················· | ···················· | ···················· | ···················· |
| Q | 8554 hCoV-19/England/MILK-ABC081/2020\|EPI_ISL_631172\|20 | ···················· | ···················· | ····················· | ···················· | ···················· | ···················· |
| Q | 8555 hCoV-19/England/ALDP-ABD2F3/2020\|EPI_ISL_631173\|20 | ···················· | ···················· | ····················· | ···················· | ···················· | ···················· |
| Q | 8556 hCoV-19/England/CAMC-AAFF48/2020\|EPI_ISL_631174\|20 | ···················· | ···················· | ····················· | ···················· | ···················· | ···················· |
| Q | 8557 hCoV-19/England/ALDP-ABD2A8/2020\|EPI_ISL_631175\|20 | ···················· | ···················· | ····················· | ···················· | ···················· | ···················· |
| Q | 8558 hCoV-19/England/MILK-ABC212/2020\|EPI_ISL_631176\|20 | ···················· | ···················· | ····················· | ···················· | ···················· | ···················· |
| Q | 8559 hCoV-19/England/CAMC-AAF740/2020\|EPI_ISL_631177\|20 | ···················· | ···················· | ····················· | ···················· | ···················· | ···················· |
| Q | 8560 hCoV-19/England/MILK-ABBB45/2020\|EPI_ISL_631178\|20 | ···················· | ···················· | ····················· | ···················· | ···················· | ···················· |
| Q | 8561 hCoV-19/England/MILK-ABBAEE/2020\|EPI_ISL_631179\|20 | ···················· | ···················· | ····················· | ···················· | ···················· | ···················· |
| Q | 8562 hCoV-19/England/CAMC-AAF555/2020\|EPI_ISL_631180\|20 | ···················· | ···················· | ····················· | ···················· | ·····T·············· | ···················· |
| Q | 8563 hCoV-19/England/CAMC-AAF810/2020\|EPI_ISL_631181\|20 | ···················· | ···················· | ····················· | ···················· | ···················· | ···················· |
| Q | 8564 hCoV-19/England/MILK-ABB8A8/2020\|EPI_ISL_631182\|20 | ···················· | ···················· | ····················· | ···················· | ···················· | ···················· |
| Q | 8565 hCoV-19/England/MILK-ABBDC7/2020\|EPI_ISL_631183\|20 | ···················· | ·A·················· | ····················· | ···················· | ···················· | ···················· |
| Q | 8566 hCoV-19/England/ALDP-AC12B0/2020\|EPI_ISL_631184\|20 | ···················· | ···················· | ····················· | ···················· | ···················· | ···················· |
| Q | 8567 hCoV-19/England/CAMC-AAF78C/2020\|EPI_ISL_631185\|20 | ···················· | ···················· | ····················· | ···················· | ···················· | ···················· |
| Q | 8568 hCoV-19/England/MILK-ABBE2E/2020\|EPI_ISL_631186\|20 | ···················· | ···················· | ····················· | ···················· | ···················· | ···················· |
| Q | 8569 hCoV-19/England/MILK-ABBF3A/2020\|EPI_ISL_631187\|20 | ···················· | ···················· | ····················· | ···················· | ···················· | ···················· |
| Q | 8570 hCoV-19/England/MILK-ABC063/2020\|EPI_ISL_631188\|20 | ···················· | ···················· | ····················· | ···················· | ···················· | ···················· |
| Q | 8571 hCoV-19/England/ALDP-AC5D06/2020\|EPI_ISL_631189\|20 | ···················· | ···················· | ····················· | ···················· | ···················· | ···················· |
| Q | 8572 hCoV-19/England/ALDP-AC11D1/2020\|EPI_ISL_631190\|20 | ···················· | ···················· | ····················· | ···················· | ···················· | ···················· |
| Q | 8573 hCoV-19/England/CAMC-AAFA29/2020\|EPI_ISL_631191\|20 | ···················· | ···················· | ····················· | ···················· | ···················· | ···················· |
| Q | 8574 hCoV-19/England/MILK-ABBD4F/2020\|EPI_ISL_631193\|20 | ···················· | ···················· | ····················· | ···················· | ···················· | ···················· |
| Q | 8575 hCoV-19/England/CAMC-AAF898/2020\|EPI_ISL_631194\|20 | ···················· | ···················· | ····················· | ···················· | ···················· | ···················· |
| Q | 8576 hCoV-19/England/MILK-ABC151/2020\|EPI_ISL_631196\|20 | ···················· | ···················· | ····················· | ···················· | ···················· | ···················· |
| Q | 8577 hCoV-19/England/MILK-ABBED3/2020\|EPI_ISL_631197\|20 | ···················· | ···················· | ····················· | ···················· | ···················· | ···················· |
| Q | 8578 hCoV-19/England/MILK-ABBD8B/2020\|EPI_ISL_631198\|20 | ···················· | ···················· | ····················· | ···················· | ···················· | ···················· |
| Q | 8579 hCoV-19/England/CAMC-AAF661/2020\|EPI_ISL_631199\|20 | ···················· | ···················· | ····················· | ···················· | ···················· | ···················· |
| Q | 8580 hCoV-19/mink/Netherlands/NB-EMC-57-4/2020\|EPI_ISL_ | ···················· | ···················· | ····················· | ···················· | ·····T·············· | ···················· |
| Q | 8581 hCoV-19/mink/Netherlands/NB-EMC-57-5/2020\|EPI_ISL_ | ···················· | ···················· | ····················· | ···················· | ·····T·············· | ···················· |
| Q | 8582 hCoV-19/Netherlands/UT-EMC-100/2020\|EPI_ISL_632460 | ···················· | ···················· | ····················· | ···················· | ···················· | ···················· |
| Q | 8583 hCoV-19/Netherlands/UT-EMC-105/2020\|EPI_ISL_632461 | ···················· | ···················· | ····················· | ···················· | ·····T·············· | ···················· |
| Q | 8584 hCoV-19/mink/Netherlands/NB-EMC-58-1/2020\|EPI_ISL_ | ···················· | ···················· | ····················· | ···················· | ·····T·············· | ···················· |
| Q | 8585 hCoV-19/mink/Netherlands/NB-EMC-58-3/2020\|EPI_ISL_ | ···················· | ···················· | ····················· | ···················· | ·····T·············· | ···················· |
| Q | 8586 hCoV-19/mink/Netherlands/NB-EMC-58-5/2020\|EPI_ISL_ | ···················· | ···················· | ····················· | ···················· | ·····T·············· | ···················· |
| Q | 8587 hCoV-19/mink/Netherlands/NB-EMC-59-2/2020\|EPI_ISL_ | ···················· | ···················· | ····················· | ···················· | ·····T·············· | ···················· |
| Q | 8588 hCoV-19/mink/Netherlands/NB-EMC-59-4/2020\|EPI_ISL_ | ···················· | ···················· | ····················· | ···················· | ·····T·············· | ···················· |
| Q | 8589 hCoV-19/mink/Netherlands/NB-EMC-59-6/2020\|EPI_ISL_ | ···················· | ···················· | ····················· | ···················· | ·····T·············· | ···················· |
| Q | 8590 hCoV-19/mink/Netherlands/NB-EMC-59-7/2020\|EPI_ISL_ | ···················· | ···················· | ····················· | ···················· | ·····T·············· | ···················· |
| Q | 8591 hCoV-19/mink/Netherlands/NB-EMC-59-8/2020\|EPI_ISL_ | ···················· | ···················· | ····················· | ···················· | ·····T·············· | ···················· |
| Q | 8592 hCoV-19/mink/Netherlands/NB-EMC-60-1/2020\|EPI_ISL_ | ···················· | ···················· | ····················· | ···················· | ·····T·············· | ···················· |
| Q | 8593 hCoV-19/mink/Netherlands/NB-EMC-60-2/2020\|EPI_ISL_ | ···················· | ···················· | ····················· | ···················· | ·····T·············· | ···················· |
| Q | 8594 hCoV-19/mink/Netherlands/NB-EMC-60-3/2020\|EPI_ISL_ | ···················· | ···················· | ····················· | ···················· | ·····T·············· | ···················· |
| Q | 8595 hCoV-19/mink/Netherlands/NB-EMC-60-4/2020\|EPI_ISL_ | ···················· | ···················· | ····················· | ···················· | ·····T·············· | ···················· |
| Q | 8596 hCoV-19/mink/Netherlands/NB-EMC-60-5/2020\|EPI_ISL_ | ···················· | ···················· | ····················· | ···················· | ·····T·············· | ···················· |
| Q | 8597 hCoV-19/mink/Netherlands/NB-EMC-61-1/2020\|EPI_ISL_ | ···················· | ···················· | ····················· | ···················· | ·····T·············· | ···················· |
| Q | 8598 hCoV-19/mink/Netherlands/NB-EMC-61-2/2020\|EPI_ISL_ | ···················· | ···················· | ····················· | ···················· | ·····T·············· | ···················· |
| Q | 8599 hCoV-19/mink/Netherlands/NB-EMC-61-3/2020\|EPI_ISL_ | ···················· | ···················· | ····················· | ···················· | ·····T·············· | ···················· |
| Q | 8600 hCoV-19/mink/Netherlands/NB-EMC-61-4/2020\|EPI_ISL_ | ···················· | ···················· | ····················· | ···················· | ·····T·············· | ···················· |
| Q | 8601 hCoV-19/Netherlands/LI-EMC-91/2020\|EPI_ISL_632479\| | ···················· | ···················· | ····················· | ···················· | ·····T·············· | ···················· |
| Q | 8602 hCoV-19/Netherlands/GE-EMC-319/2020\|EPI_ISL_632480 | ···················· | ···················· | ····················· | ···················· | ···················· | ···················· |
| Q | 8603 hCoV-19/Netherlands/GE-EMC-322/2020\|EPI_ISL_632481 | ···················· | ···················· | ····················· | ···················· | ···················· | ···················· |
| Q | 8604 hCoV-19/Netherlands/GE-EMC-334/2020\|EPI_ISL_632482 | ···················· | ·A·················· | ····················· | ···················· | ···················· | ··A················· |
| Q | 8605 hCoV-19/mink/Netherlands/NB-EMC-63-1/2020\|EPI_ISL_ | ···················· | ···················· | ····················· | ···················· | ·····T·············· | ···················· |
| Q | 8606 hCoV-19/mink/Netherlands/NB-EMC-63-4/2020\|EPI_ISL_ | ···················· | ···················· | ····················· | ···················· | ·····T·············· | ···················· |
| Q | 8607 hCoV-19/mink/Netherlands/NB-EMC-63-5/2020\|EPI_ISL_ | ···················· | ···················· | ····················· | ···················· | ·····T·············· | ···················· |
| Q | 8608 hCoV-19/mink/Netherlands/NB-EMC-64-1/2020\|EPI_ISL_ | ···················· | ···················· | ····················· | ···················· | ·····T·············· | ···················· |
| Q | 8609 hCoV-19/mink/Netherlands/NB-EMC-64-3/2020\|EPI_ISL_ | ···················· | ···················· | ····················· | ···················· | ·····T·············· | ···················· |
| Q | 8610 hCoV-19/mink/Netherlands/NB-EMC-64-7/2020\|EPI_ISL_ | ···················· | ···················· | ····················· | ···················· | ·····T·············· | ···················· |
| Q | 8611 hCoV-19/mink/Netherlands/NB-EMC-65-1/2020\|EPI_ISL_ | ···················· | ···················· | ····················· | ···················· | ·····T·············· | ···················· |
| Q | 8612 hCoV-19/mink/Netherlands/NB-EMC-65-3/2020\|EPI_ISL_ | ···················· | ···················· | ····················· | ···················· | ·····T·············· | ···················· |
| Q | 8613 hCoV-19/mink/Netherlands/NB-EMC-65-10/2020\|EPI_ISL | ···················· | ···················· | ····················· | ···················· | ·····T·············· | ···················· |
| Q | 8614 hCoV-19/mink/Netherlands/NB-EMC/2020\|EPI_ISL_63249 | ···················· | ···················· | ····················· | ···················· | ·····T·············· | ···················· |
| Q | 8615 hCoV-19/Netherlands/ZH-EMC-652/2020\|EPI_ISL_632494 | ···················· | ···················· | ····················· | ···················· | ···················· | ···················· |
| Q | 8616 hCoV-19/Netherlands/ZH-EMC-655/2020\|EPI_ISL_632495 | ···················· | ···················· | ····················· | ···················· | ·····T·············· | ···················· |
| Q | 8617 hCoV-19/Netherlands/UT-EMC-92/2020\|EPI_ISL_632496\| | ···················· | ···················· | ····················· | ···················· | ···················· | ···················· |
| Q | 8618 hCoV-19/Netherlands/UT-EMC-93/2020\|EPI_ISL_632497\| | ···················· | ···················· | ····················· | ···················· | ···················· | ···················· |
| Q | 8619 hCoV-19/Netherlands/UT-EMC-94/2020\|EPI_ISL_632498\| | ···················· | ···················· | ····················· | ···················· | ···················· | ···················· |

| | | | | | | | | |
|---|---|---|---|---|---|---|---|---|
| Q | 8620 | hCoV-19/Netherlands/UT-EMC-95/2020\|EPI_ISL_632499\| | ···················· | ···················· | ····················· | ···················· | ···················· | ···················· |
| Q | 8621 | hCoV-19/England/QEUH-AD1B76/2020\|EPI_ISL_633723\|20 | ···················· | ···················· | ····················· | ···················· | ···················· | ···················· |
| Q | 8622 | hCoV-19/England/QEUH-AD59C3/2020\|EPI_ISL_633724\|20 | ···················· | ···················· | ····················· | ···················· | ···················· | ···················· |
| Q | 8623 | hCoV-19/England/QEUH-AD3145/2020\|EPI_ISL_633725\|20 | ···················· | ···················· | ····················· | ···················· | ···················· | ···················· |
| Q | 8624 | hCoV-19/England/QEUH-AD0B86/2020\|EPI_ISL_633726\|20 | ···················· | ···················· | ····················· | ···················· | ···················· | ···················· |
| Q | 8625 | hCoV-19/England/QEUH-AD1129/2020\|EPI_ISL_633727\|20 | ···················· | ···················· | ····················· | ···················· | ···················· | ···················· |
| Q | 8626 | hCoV-19/England/QEUH-AD28AB/2020\|EPI_ISL_633728\|20 | ···················· | ·A·················· | ····················· | ···················· | ···················· | ··A················· |
| Q | 8627 | hCoV-19/England/QEUH-AD1BD0/2020\|EPI_ISL_633729\|20 | ···················· | ···················· | ····················· | ···················· | ···················· | ···················· |
| Q | 8628 | hCoV-19/England/QEUH-AD0B3B/2020\|EPI_ISL_633731\|20 | ···················· | ···················· | ····················· | ···················· | ···················· | ···················· |
| Q | 8629 | hCoV-19/England/QEUH-AD32F7/2020\|EPI_ISL_633732\|20 | ···················· | ···················· | ····················· | ···················· | ···················· | ···················· |
| Q | 8630 | hCoV-19/England/QEUH-AD54B3/2020\|EPI_ISL_633733\|20 | ···················· | ···················· | ····················· | ···················· | ···················· | ···················· |
| Q | 8631 | hCoV-19/England/QEUH-AD3233/2020\|EPI_ISL_633789\|20 | ···················· | ···················· | ····················· | ···················· | ···················· | ···················· |
| Q | 8632 | hCoV-19/England/QEUH-AD10D1/2020\|EPI_ISL_633790\|20 | ···················· | ···················· | ····················· | ···················· | ···················· | ···················· |
| Q | 8633 | hCoV-19/England/QEUH-AD1B3A/2020\|EPI_ISL_633791\|20 | ···················· | ···················· | ····················· | ···················· | ···················· | ···················· |
| Q | 8634 | hCoV-19/England/QEUH-AD14D5/2020\|EPI_ISL_633792\|20 | ···················· | ···················· | ····················· | ···················· | ···················· | ···················· |
| Q | 8635 | hCoV-19/England/QEUH-AD11ED/2020\|EPI_ISL_633793\|20 | ···················· | ···················· | ····················· | ···················· | ···················· | ···················· |
| Q | 8636 | hCoV-19/England/QEUH-AD386E/2020\|EPI_ISL_633795\|20 | ···················· | ···················· | ····················· | ···················· | ···················· | ···················· |
| Q | 8637 | hCoV-19/England/QEUH-AD10A4/2020\|EPI_ISL_633796\|20 | ···················· | ···················· | ····················· | ···················· | ···················· | ···················· |
| Q | 8638 | hCoV-19/England/QEUH-AD3691/2020\|EPI_ISL_633797\|20 | ···················· | ···················· | ····················· | ···················· | ···················· | ···················· |
| Q | 8639 | hCoV-19/England/QEUH-AD089E/2020\|EPI_ISL_633798\|20 | ···················· | ···················· | ····················· | ···················· | ···················· | ···················· |
| R | 1 | hCoV-19/Australia/VIC13200/2020\|EPI_ISL_641302\|202 | ···················· | ···················· | ····················· | ···················· | ···················· | ···················· |
| R | 2 | hCoV-19/Australia/VIC16391/2020\|EPI_ISL_641303\|202 | ···················· | ···················· | ····················· | ···················· | ···················· | ···················· |
| R | 3 | hCoV-19/Australia/VIC15937/2020\|EPI_ISL_641307\|202 | ···················· | ···················· | ····················· | ···················· | ···················· | ···················· |
| R | 4 | hCoV-19/Australia/VIC16516/2020\|EPI_ISL_641308\|202 | ···················· | ···················· | ····················· | ···················· | ···················· | ···················· |
| R | 5 | hCoV-19/Australia/QLD1159/2020\|EPI_ISL_641309\|2020 | ···················· | ···················· | ····················· | ···················· | ···················· | ···················· |
| R | 6 | hCoV-19/Australia/QLD1161/2020\|EPI_ISL_641310\|2020 | ···················· | ···················· | ····················· | ···················· | ···················· | ···················· |
| R | 7 | hCoV-19/England/QEUH-B126A4/2020\|EPI_ISL_642611\|20 | ···················· | ·····C·············· | ····················· | ···················· | ···················· | ···················· |
| R | 8 | hCoV-19/England/QEUH-B144A8/2020\|EPI_ISL_642612\|20 | ···················· | ···················· | ····················· | ···················· | ···················· | ···················· |
| R | 9 | hCoV-19/England/QEUH-B1005D/2020\|EPI_ISL_642613\|20 | ···················· | ···················· | ····················· | ···················· | ···················· | ···················· |
| R | 10 | hCoV-19/England/QEUH-B108FB/2020\|EPI_ISL_642614\|20 | ···················· | ···················· | ····················· | ···················· | ···················· | ···················· |
| R | 11 | hCoV-19/England/QEUH-B12AD5/2020\|EPI_ISL_642615\|20 | ···················· | ···················· | ····················· | ···················· | ···················· | ···················· |
| R | 12 | hCoV-19/England/QEUH-B1264A/2020\|EPI_ISL_642616\|20 | ···················· | ···················· | ····················· | ···················· | ···················· | ···················· |
| R | 13 | hCoV-19/England/QEUH-B10011/2020\|EPI_ISL_642617\|20 | ···················· | ···················· | ····················· | ···················· | ···················· | ···················· |
| R | 14 | hCoV-19/England/QEUH-B143F6/2020\|EPI_ISL_642618\|20 | ···················· | ···················· | ····················· | ···················· | ···················· | ···················· |
| R | 15 | hCoV-19/England/QEUH-B11308/2020\|EPI_ISL_642619\|20 | ···················· | ···················· | ····················· | ···················· | ···················· | ···················· |
| R | 16 | hCoV-19/England/QEUH-B145F0/2020\|EPI_ISL_642620\|20 | ···················· | ···················· | ····················· | ···················· | ···················· | ···················· |
| R | 17 | hCoV-19/England/QEUH-B12677/2020\|EPI_ISL_642621\|20 | ···················· | ···················· | ····················· | ···················· | ···················· | ···················· |
| R | 18 | hCoV-19/England/QEUH-B1003F/2020\|EPI_ISL_642622\|20 | ···················· | ···················· | ····················· | ···················· | ···················· | ···················· |
| R | 19 | hCoV-19/England/QEUH-B113DB/2020\|EPI_ISL_642623\|20 | ···················· | ···················· | ····················· | ···················· | ···················· | ···················· |
| R | 20 | hCoV-19/England/QEUH-B12BD2/2020\|EPI_ISL_642626\|20 | ···················· | ···················· | ····················· | ···················· | ···················· | ···················· |
| R | 21 | hCoV-19/England/QEUH-B12BC3/2020\|EPI_ISL_642628\|20 | ···················· | ·A·················· | ····················· | ···················· | ···················· | ···················· |
| R | 22 | hCoV-19/England/QEUH-B0ECE5/2020\|EPI_ISL_642629\|20 | ···················· | ···················· | ····················· | ···················· | ···················· | ···················· |
| R | 23 | hCoV-19/England/QEUH-B145C3/2020\|EPI_ISL_642633\|20 | ···················· | ···················· | ····················· | ···················· | ···················· | ···················· |
| R | 24 | hCoV-19/England/QEUH-B11FD7/2020\|EPI_ISL_642634\|20 | ···················· | ···················· | ····················· | ···················· | ···················· | ···················· |
| R | 25 | hCoV-19/England/QEUH-B129C9/2020\|EPI_ISL_642635\|20 | ···················· | ···················· | ····················· | ···················· | ···················· | ···················· |
| R | 26 | hCoV-19/England/QEUH-B113CC/2020\|EPI_ISL_642636\|20 | ···················· | ···················· | ····················· | ···················· | ·····T·············· | ···················· |
| R | 27 | hCoV-19/England/QEUH-B12E24/2020\|EPI_ISL_642637\|20 | ···················· | ···················· | ····················· | ···················· | ···················· | ···················· |
| R | 28 | hCoV-19/England/QEUH-B11FC8/2020\|EPI_ISL_642638\|20 | ···················· | ···················· | ····················· | ···················· | ···················· | ···················· |
| R | 29 | hCoV-19/England/QEUH-B14411/2020\|EPI_ISL_642639\|20 | ···················· | ···················· | ····················· | ···················· | ···················· | ···················· |
| R | 30 | hCoV-19/England/QEUH-B12B0F/2020\|EPI_ISL_642640\|20 | ···················· | ···················· | ····················· | ···················· | ···················· | ···················· |
| R | 31 | hCoV-19/England/QEUH-B12501/2020\|EPI_ISL_642641\|20 | ···················· | ···················· | ····················· | ···················· | ···················· | ···················· |
| R | 32 | hCoV-19/England/QEUH-B112CF/2020\|EPI_ISL_642642\|20 | ···················· | ···················· | ····················· | ···················· | ···················· | ···················· |
| R | 33 | hCoV-19/England/QEUH-B10099/2020\|EPI_ISL_642643\|20 | ···················· | ···················· | ····················· | ···················· | ···················· | ···················· |
| R | 34 | hCoV-19/England/QEUH-B12A99/2020\|EPI_ISL_642644\|20 | ···················· | ···················· | ····················· | ···················· | ···················· | ···················· |
| R | 35 | hCoV-19/England/QEUH-B12E60/2020\|EPI_ISL_642645\|20 | ···················· | ···················· | ····················· | ···················· | ···················· | ···················· |
| R | 36 | hCoV-19/England/QEUH-B143AB/2020\|EPI_ISL_642646\|20 | ···················· | ···················· | ····················· | ···················· | ···················· | ···················· |
| R | 37 | hCoV-19/England/QEUH-B11371/2020\|EPI_ISL_642648\|20 | ···················· | ···················· | ····················· | ···················· | ···················· | ···················· |
| R | 38 | hCoV-19/England/QEUH-B145D2/2020\|EPI_ISL_642649\|20 | ···················· | ···················· | ····················· | ···················· | ···················· | ···················· |
| R | 39 | hCoV-19/England/QEUH-B124AA/2020\|EPI_ISL_642650\|20 | ···················· | ···················· | ····················· | ···················· | ···················· | ···················· |
| R | 40 | hCoV-19/England/QEUH-B12BE1/2020\|EPI_ISL_642651\|20 | ···················· | ···················· | ····················· | ···················· | ···················· | ···················· |
| R | 41 | hCoV-19/England/QEUH-B125C5/2020\|EPI_ISL_642652\|20 | ···················· | ···················· | ····················· | ···················· | ···················· | ···················· |
| R | 42 | hCoV-19/England/QEUH-B12370/2020\|EPI_ISL_642654\|20 | ···················· | ···················· | ····················· | ···················· | ···················· | ···················· |
| R | 43 | hCoV-19/England/CAMC-B09303/2020\|EPI_ISL_643942\|20 | ···················· | ·A·················· | ····················· | ···················· | ···················· | ··A················· |
| R | 44 | hCoV-19/England/CAMC-B08708/2020\|EPI_ISL_643943\|20 | ···················· | ···················· | ····················· | ···················· | ···················· | ···················· |
| R | 45 | hCoV-19/England/CAMC-B07129/2020\|EPI_ISL_643944\|20 | ···················· | ···················· | ····················· | ···················· | ···················· | ···················· |
| R | 46 | hCoV-19/England/CAMC-B08B1B/2020\|EPI_ISL_643945\|20 | ···················· | ···················· | ····················· | ···················· | ···················· | ···················· |
| R | 47 | hCoV-19/England/CAMC-B074D5/2020\|EPI_ISL_643946\|20 | ···················· | ···················· | ····················· | ···················· | ···················· | ···················· |
| R | 48 | hCoV-19/England/CAMC-B07165/2020\|EPI_ISL_643947\|20 | ···················· | ···················· | ····················· | ···················· | ···················· | ···················· |
| R | 49 | hCoV-19/England/CAMC-B08638/2020\|EPI_ISL_643948\|20 | ···················· | ···················· | ····················· | ···················· | ···················· | ···················· |
| R | 50 | hCoV-19/England/CAMC-B08753/2020\|EPI_ISL_643949\|20 | ···················· | ···················· | ····················· | ···················· | ···················· | ···················· |
| R | 51 | hCoV-19/England/CAMC-B086ED/2020\|EPI_ISL_643950\|20 | ···················· | ···················· | ····················· | ···················· | ···················· | ···················· |
| R | 52 | hCoV-19/England/CAMC-B078AC/2020\|EPI_ISL_643951\|20 | ···················· | ···················· | ····················· | ···················· | ···················· | ···················· |
| R | 53 | hCoV-19/England/CAMC-B0828F/2020\|EPI_ISL_643952\|20 | ···················· | ···················· | ····················· | ···················· | ···················· | ···················· |
| R | 54 | hCoV-19/England/CAMC-B072F9/2020\|EPI_ISL_643953\|20 | ···················· | ···················· | ····················· | ···················· | ···················· | ···················· |
| R | 55 | hCoV-19/England/CAMC-B0886F/2020\|EPI_ISL_643954\|20 | ···················· | ···················· | ····················· | ···················· | ···················· | ···················· |
| R | 56 | hCoV-19/England/CAMC-B0927F/2020\|EPI_ISL_643958\|20 | ···················· | ···················· | ····················· | ···················· | ···················· | ···················· |
| R | 57 | hCoV-19/England/CAMC-B07684/2020\|EPI_ISL_643959\|20 | ···················· | ···················· | ····················· | ···················· | ···················· | ···················· |
| R | 58 | hCoV-19/England/CAMC-B070C2/2020\|EPI_ISL_643964\|20 | ···················· | ···················· | ····················· | ···················· | ···················· | ···················· |
| R | 59 | hCoV-19/England/CAMC-B08BDF/2020\|EPI_ISL_643965\|20 | ···················· | ···················· | ····················· | ···················· | ···················· | ···················· |
| R | 60 | hCoV-19/England/CAMC-B08C54/2020\|EPI_ISL_643966\|20 | ···················· | ···················· | ····················· | ···················· | ···················· | ···················· |

| | | | | | | | |
|---|---|---|---|---|---|---|---|
| R | 61 hCoV-19/England/CAMC-B07EE6/2020\|EPI_ISL_643968\|20 | ···················· | ···················· | ····················· | ···················· | ···················· | ···················· |
| R | 62 hCoV-19/England/CAMC-B07305/2020\|EPI_ISL_643970\|20 | ···················· | ···················· | ····················· | ···················· | ···················· | ···················· |
| R | 63 hCoV-19/England/CAMC-B08AE1/2020\|EPI_ISL_643972\|20 | ···················· | ···················· | ····················· | ···················· | ···················· | ···················· |
| R | 64 hCoV-19/England/CAMC-B09233/2020\|EPI_ISL_643973\|20 | ···················· | ···················· | ····················· | ···················· | ···················· | ···················· |
| R | 65 hCoV-19/England/CAMC-B07280/2020\|EPI_ISL_643974\|20 | ···················· | ···················· | ····················· | ···················· | ···················· | ···················· |
| R | 66 hCoV-19/England/CAMC-B0737E/2020\|EPI_ISL_643975\|20 | ···················· | ···················· | ····················· | ···················· | ···················· | ···················· |
| R | 67 hCoV-19/England/CAMC-B08647/2020\|EPI_ISL_643976\|20 | ···················· | ···················· | ····················· | ···················· | ···················· | ···················· |
| R | 68 hCoV-19/England/CAMC-B07ED7/2020\|EPI_ISL_643977\|20 | ···················· | ···················· | ····················· | ···················· | ·····T·············· | ···················· |
| R | 69 hCoV-19/England/CAMC-B07F2F/2020\|EPI_ISL_643978\|20 | ···················· | ···················· | ····················· | ···················· | ···················· | ···················· |
| R | 70 hCoV-19/England/CAMC-B074E4/2020\|EPI_ISL_643979\|20 | ···················· | ···················· | ····················· | ···················· | ·····T·············· | ···················· |
| R | 71 hCoV-19/England/CAMC-B0788E/2020\|EPI_ISL_643980\|20 | ···················· | ···················· | ····················· | ···················· | ···················· | ···················· |
| R | 72 hCoV-19/England/CAMC-B08726/2020\|EPI_ISL_643981\|20 | ···················· | ···················· | ····················· | ···················· | ···················· | ···················· |
| R | 73 hCoV-19/England/CAMC-B076A2/2020\|EPI_ISL_643982\|20 | ···················· | ···················· | ····················· | ···················· | ···················· | ···················· |
| R | 74 hCoV-19/England/MILK-ACD1B7/2020\|EPI_ISL_643984\|20 | ···················· | ···················· | ····················· | ···················· | ···················· | ···················· |
| R | 75 hCoV-19/England/ALDP-B1AC5F/2020\|EPI_ISL_646100\|20 | ···················· | ···················· | ····················· | ···················· | ···················· | ···················· |
| R | 76 hCoV-19/England/CAMC-B21D52/2020\|EPI_ISL_646101\|20 | ···················· | ···················· | ····················· | ···················· | ···················· | ···················· |
| R | 77 hCoV-19/England/CAMC-B2210A/2020\|EPI_ISL_646102\|20 | ···················· | ···················· | ····················· | ···················· | ···················· | ···················· |
| R | 78 hCoV-19/England/ALDP-B1C121/2020\|EPI_ISL_646103\|20 | ···················· | ···················· | ····················· | ···················· | ···················· | ···················· |
| R | 79 hCoV-19/England/ALDP-B1ADE3/2020\|EPI_ISL_646104\|20 | ···················· | ···················· | ····················· | ···················· | ···················· | ···················· |
| R | 80 hCoV-19/England/CAMC-B22586/2020\|EPI_ISL_646105\|20 | ···················· | ···················· | ····················· | ···················· | ···················· | ···················· |
| R | 81 hCoV-19/England/ALDP-B1DBAA/2020\|EPI_ISL_646106\|20 | ···················· | ···················· | ····················· | ···················· | ···················· | ···················· |
| R | 82 hCoV-19/England/ALDP-B1A2A8/2020\|EPI_ISL_646107\|20 | ···················· | ···················· | ····················· | ···················· | ···················· | ···················· |
| R | 83 hCoV-19/England/ALDP-B1DAE9/2020\|EPI_ISL_646108\|20 | ···················· | ···················· | ····················· | ···················· | ···················· | ···················· |
| R | 84 hCoV-19/England/ALDP-B1F7B3/2020\|EPI_ISL_646109\|20 | ···················· | ···················· | ····················· | ···················· | ···················· | ···················· |
| R | 85 hCoV-19/England/ALDP-B1C464/2020\|EPI_ISL_646110\|20 | ···················· | ···················· | ····················· | ···················· | ···················· | ···················· |
| R | 86 hCoV-19/England/ALDP-B1A35A/2020\|EPI_ISL_646111\|20 | ···················· | ···················· | ····················· | ···················· | ···················· | ···················· |
| R | 87 hCoV-19/England/ALDP-B1A2F3/2020\|EPI_ISL_646112\|20 | ···················· | ···················· | ····················· | ···················· | ···················· | ···················· |
| R | 88 hCoV-19/England/ALDP-B1D603/2020\|EPI_ISL_646113\|20 | ···················· | ···················· | ····················· | ···················· | ···················· | ···················· |
| R | 89 hCoV-19/England/ALDP-B1A40C/2020\|EPI_ISL_646114\|20 | ···················· | ···················· | ····················· | ···················· | ···················· | ···················· |
| R | 90 hCoV-19/England/CAMC-B220B2/2020\|EPI_ISL_646115\|20 | ···················· | ···················· | ····················· | ···················· | ···················· | ···················· |
| R | 91 hCoV-19/England/ALDP-B1AD5C/2020\|EPI_ISL_646116\|20 | ···················· | ···················· | ····················· | ···················· | ···················· | ···················· |
| R | 92 hCoV-19/England/ALDP-B1A3D2/2020\|EPI_ISL_646117\|20 | ···················· | ···················· | ····················· | ···················· | ···················· | ···················· |
| R | 93 hCoV-19/England/ALDP-B1A703/2020\|EPI_ISL_646118\|20 | ···················· | ···················· | ····················· | ···················· | ···················· | ···················· |
| R | 94 hCoV-19/England/ALDP-B1A387/2020\|EPI_ISL_646119\|20 | ···················· | ···················· | ····················· | ···················· | ···················· | ···················· |
| R | 95 hCoV-19/England/ALDP-B1DB22/2020\|EPI_ISL_646120\|20 | ···················· | ···················· | ····················· | ···················· | ···················· | ···················· |
| R | 96 hCoV-19/England/ALDP-B1D5BB/2020\|EPI_ISL_646121\|20 | ···················· | ···················· | ····················· | ···················· | ···················· | ···················· |
| R | 97 hCoV-19/England/CAMC-B21FB6/2020\|EPI_ISL_646122\|20 | ···················· | ·A······A··········· | ····················· | ···················· | ···················· | ···················· |
| R | 98 hCoV-19/England/ALDP-B1A28A/2020\|EPI_ISL_646123\|20 | ···················· | ···················· | ····················· | ···················· | ···················· | ···················· |
| R | 99 hCoV-19/England/ALDP-B1F5C8/2020\|EPI_ISL_646124\|20 | ···················· | ···················· | ····················· | ···················· | ···················· | ···················· |
| R | 100 hCoV-19/England/ALDP-B1FA05/2020\|EPI_ISL_646125\|20 | ···················· | ···················· | ····················· | ···················· | ···················· | ···················· |
| R | 101 hCoV-19/England/ALDP-B1B83C/2020\|EPI_ISL_646126\|20 | ···················· | ···················· | ····················· | ···················· | ···················· | ···················· |
| R | 102 hCoV-19/England/ALDP-B1AD10/2020\|EPI_ISL_646127\|20 | ···················· | ···················· | ····················· | ···················· | ···················· | ···················· |
| R | 103 hCoV-19/England/CAMC-B22216/2020\|EPI_ISL_646128\|20 | ···················· | ···················· | ····················· | ···················· | ···················· | ···················· |
| R | 104 hCoV-19/England/CAMC-B2242F/2020\|EPI_ISL_646129\|20 | ···················· | ···················· | ····················· | ···················· | ·····T·············· | ···················· |
| R | 105 hCoV-19/England/ALDP-B1F62F/2020\|EPI_ISL_646130\|20 | ···················· | ···················· | ····················· | ···················· | ···················· | ···················· |
| R | 106 hCoV-19/England/CAMC-B222BC/2020\|EPI_ISL_646131\|20 | ···················· | ···················· | ····················· | ···················· | ···T················ | ···················· |
| R | 107 hCoV-19/England/ALDP-B1DB04/2020\|EPI_ISL_646132\|20 | ···················· | ···················· | ····················· | ···················· | ···················· | ···················· |
| R | 108 hCoV-19/England/ALDP-B1F59B/2020\|EPI_ISL_646133\|20 | ···················· | ···················· | ····················· | ···················· | ···················· | ···················· |
| R | 109 hCoV-19/England/ALDP-B1F8B0/2020\|EPI_ISL_646134\|20 | ···················· | ···················· | ····················· | ···················· | ···················· | ···················· |
| R | 110 hCoV-19/England/CAMC-B22401/2020\|EPI_ISL_646135\|20 | ···················· | ···················· | ····················· | ···················· | ···················· | ···················· |
| R | 111 hCoV-19/England/CAMC-B22331/2020\|EPI_ISL_646136\|20 | ···················· | ···················· | ····················· | ···················· | ···················· | ···················· |
| R | 112 hCoV-19/England/ALDP-B1DAAD/2020\|EPI_ISL_646137\|20 | ···················· | ···················· | ····················· | ···················· | ···················· | ···················· |
| R | 113 hCoV-19/England/ALDP-B1A3E1/2020\|EPI_ISL_646138\|20 | ···················· | ···················· | ····················· | ···················· | ···················· | ···················· |
| R | 114 hCoV-19/England/CAMC-B2200D/2020\|EPI_ISL_646139\|20 | ···················· | ···················· | ····················· | ···················· | ···················· | ···················· |
| R | 115 hCoV-19/England/QEUH-B31B37/2020\|EPI_ISL_646140\|20 | ···················· | ···················· | ····················· | ···················· | ···················· | ···················· |
| R | 116 hCoV-19/England/QEUH-B31D22/2020\|EPI_ISL_646141\|20 | ···················· | ···················· | ····················· | ···················· | ···················· | ···················· |
| R | 117 hCoV-19/England/QEUH-B2E1C2/2020\|EPI_ISL_646142\|20 | ···················· | ···················· | ····················· | ···················· | ···················· | ···················· |
| R | 118 hCoV-19/England/QEUH-B310DE/2020\|EPI_ISL_646143\|20 | ···················· | ···················· | ····················· | ···················· | ···················· | ···················· |
| R | 119 hCoV-19/England/QEUH-B31C8F/2020\|EPI_ISL_646144\|20 | ···················· | ···················· | ····················· | ···················· | ···················· | ···················· |
| R | 120 hCoV-19/England/QEUH-B2F756/2020\|EPI_ISL_646145\|20 | ···················· | ···················· | ····················· | ···················· | ···················· | ···················· |
| R | 121 hCoV-19/England/QEUH-B31A0D/2020\|EPI_ISL_646146\|20 | ···················· | ···················· | ····················· | ···················· | ·····T·············· | ···················· |
| R | 122 hCoV-19/England/QEUH-B31B64/2020\|EPI_ISL_646147\|20 | ···················· | ···················· | ····················· | ···················· | ···················· | ···················· |
| R | 123 hCoV-19/England/ALDP-B240DE/2020\|EPI_ISL_646148\|20 | ···················· | ···················· | ····················· | ···················· | ···················· | ···················· |
| R | 124 hCoV-19/England/QEUH-B31D13/2020\|EPI_ISL_646149\|20 | ···················· | ···················· | ····················· | ···················· | ···················· | ···················· |
| R | 125 hCoV-19/England/ALDP-B23C17/2020\|EPI_ISL_646150\|20 | ···················· | ···················· | ····················· | ···················· | ···················· | ···················· |
| R | 126 hCoV-19/England/QEUH-B31F68/2020\|EPI_ISL_646151\|20 | ···················· | ···················· | ····················· | ···················· | ···················· | ···················· |
| R | 127 hCoV-19/England/QEUH-B2E1A4/2020\|EPI_ISL_646152\|20 | ···················· | ·A·················· | ····················· | ···················· | ···················· | ···················· |
| R | 128 hCoV-19/England/QEUH-B2E02F/2020\|EPI_ISL_646153\|20 | ···················· | ···················· | ····················· | ···················· | ···················· | ···················· |
| R | 129 hCoV-19/England/QEUH-B2E21A/2020\|EPI_ISL_646154\|20 | ···················· | ···················· | ····················· | ···················· | ···················· | ···················· |
| R | 130 hCoV-19/England/ALDP-B23EE4/2020\|EPI_ISL_646155\|20 | ···················· | ···················· | ····················· | ···················· | ···················· | ···················· |
| R | 131 hCoV-19/England/ALDP-B23B74/2020\|EPI_ISL_646156\|20 | ···················· | ···················· | ····················· | ···················· | ···················· | ···················· |
| R | 132 hCoV-19/England/ALDP-B23C80/2020\|EPI_ISL_646157\|20 | ···················· | ···················· | ····················· | ···················· | ···················· | ···················· |
| R | 133 hCoV-19/England/QEUH-B31FD1/2020\|EPI_ISL_646158\|20 | ···················· | ···················· | ····················· | ···················· | ···················· | ···················· |
| R | 134 hCoV-19/England/QEUH-B2F808/2020\|EPI_ISL_646159\|20 | ···················· | ···················· | ····················· | ···················· | ···················· | ···················· |
| R | 135 hCoV-19/England/QEUH-B31DD7/2020\|EPI_ISL_646160\|20 | ···················· | ···················· | ····················· | ···················· | ···················· | ···················· |
| R | 136 hCoV-19/England/QEUH-B31E89/2020\|EPI_ISL_646161\|20 | ···················· | ···················· | ····················· | ···················· | ···················· | ···················· |
| R | 137 hCoV-19/England/ALDP-B23DC9/2020\|EPI_ISL_646162\|20 | ···················· | ···················· | ····················· | ···················· | ···················· | ···················· |
| R | 138 hCoV-19/England/QEUH-B31CCB/2020\|EPI_ISL_646163\|20 | ···················· | ···················· | ····················· | ···················· | ···················· | ···················· |
| R | 139 hCoV-19/England/QEUH-B2E14A/2020\|EPI_ISL_646164\|20 | ···················· | ···················· | ····················· | ···················· | ·····T·············· | ···················· |
| R | 140 hCoV-19/England/ALDP-B23F5A/2020\|EPI_ISL_646166\|20 | ···················· | ·A·················· | ····················· | ···················· | ···················· | ···················· |

R 141 hCoV-19/England/QEUH-B319A6/2020|EPI_ISL_646167|20 ···················· ···················· ···················· ···················· ···················· ····················
R 142 hCoV-19/England/QEUH-B2E49C/2020|EPI_ISL_646168|20 ···················· ···················· ···················· ···················· ···················· ····················
R 143 hCoV-19/England/ALDP-B24162/2020|EPI_ISL_646169|20 ···················· ···················· ···················· ···················· ···················· ····················
R 144 hCoV-19/England/ALDP-B23ED5/2020|EPI_ISL_646170|20 ···················· ···················· ···················· ···················· ···················· ····················
R 145 hCoV-19/England/QEUH-B3195B/2020|EPI_ISL_646171|20 ···················· ···················· ···················· ···················· ···················· ····················
R 146 hCoV-19/England/ALDP-B23BA1/2020|EPI_ISL_646173|20 ···················· ···················· ···················· ···················· ······T············· ····················
R 147 hCoV-19/England/QEUH-B2E502/2020|EPI_ISL_646174|20 ···················· ···················· ···················· ···················· ···················· ····················
R 148 hCoV-19/England/QEUH-B32073/2020|EPI_ISL_646175|20 ···················· ···················· ···················· ···················· ···················· ····················
R 149 hCoV-19/England/QEUH-B320BF/2020|EPI_ISL_646176|20 ···················· ···················· ···················· ···················· ···················· ····················
R 150 hCoV-19/England/QEUH-B31B82/2020|EPI_ISL_646177|20 ···················· ···················· ···················· ···················· ···················· ····················
R 151 hCoV-19/England/QEUH-B31EE3/2020|EPI_ISL_646178|20 ···················· ···················· ···················· ···················· ···················· ····················
R 152 hCoV-19/England/QEUH-B2E511/2020|EPI_ISL_646179|20 ···················· ···················· ···················· ···················· ···················· ····················
R 153 hCoV-19/England/CAMC-B22D9D/2020|EPI_ISL_647429|20 ···················· ·A·················· ···················· ···················· ···················· ···A················
R 154 hCoV-19/England/CAMC-B22191/2020|EPI_ISL_647430|20 ···················· ···················· ···················· ···················· ···················· ····················
R 155 hCoV-19/England/CAMC-B22173/2020|EPI_ISL_647431|20 ···················· ···················· ···················· ···················· ···················· ····················
R 156 hCoV-19/England/CAMC-B2147B/2020|EPI_ISL_647432|20 ···················· ···················· ···················· ···················· ···················· ····················
R 157 hCoV-19/England/CAMC-B21657/2020|EPI_ISL_647433|20 ···················· ···················· ···················· ···················· ···················· ····················
R 158 hCoV-19/England/CAMC-B22F3D/2020|EPI_ISL_647434|20 ···················· ···················· ···················· ···················· ···················· ····················
R 159 hCoV-19/England/CAMC-B21736/2020|EPI_ISL_647435|20 ···················· ·A·················· ···················· ···················· ···················· ···A················
R 160 hCoV-19/England/CAMC-B21860/2020|EPI_ISL_647436|20 ···················· ···················· ···················· ···················· ···················· ····················
R 161 hCoV-19/England/CAMC-B215E1/2020|EPI_ISL_647437|20 ···················· ···················· ···················· ···················· ···················· ····················
R 162 hCoV-19/England/CAMC-B2246B/2020|EPI_ISL_647438|20 ···················· ···················· ···················· ···················· ······T············· ····················
R 163 hCoV-19/England/CAMC-B22DD9/2020|EPI_ISL_647439|20 ···················· ···················· ···················· ···················· ···················· ····················
R 164 hCoV-19/England/CAMC-B2247A/2020|EPI_ISL_647440|20 ···················· ···················· ···················· ···················· ···················· ····················
R 165 hCoV-19/England/CAMC-B22A69/2020|EPI_ISL_647441|20 ···················· ···················· ···················· ···················· ···················· ····················
R 166 hCoV-19/England/CAMC-B2148A/2020|EPI_ISL_647442|20 ···················· ···················· ···················· ···················· ···················· ····················
R 167 hCoV-19/England/CAMC-B216FD/2020|EPI_ISL_647443|20 ···················· ·A·················· ···················· ···················· ···················· ···A················
R 168 hCoV-19/England/CAMC-B21499/2020|EPI_ISL_647444|20 ···················· ···················· ···················· ···················· ···················· ····················
R 169 hCoV-19/England/CAMC-B22753/2020|EPI_ISL_647446|20 ···················· ···················· ···················· ···················· ···················· ····················
R 170 hCoV-19/England/CAMC-B22A2D/2020|EPI_ISL_647447|20 ···················· ···················· ···················· ···················· ···················· ····················
R 171 hCoV-19/England/CAMC-B224F2/2020|EPI_ISL_647448|20 ···················· ···················· ···················· ···················· ···················· ····················
R 172 hCoV-19/England/CAMC-B226ED/2020|EPI_ISL_647449|20 ···················· ···················· ···················· ···················· ···················· ····················
R 173 hCoV-19/England/CAMC-B22C09/2020|EPI_ISL_647450|20 ···················· ···················· ···················· ···················· ···················· ····················
R 174 hCoV-19/England/CAMC-B21851/2020|EPI_ISL_647451|20 ···················· ···················· ···················· ···················· ···················· ····················
R 175 hCoV-19/England/CAMC-B22B48/2020|EPI_ISL_647453|20 ···················· ···················· ···················· ···················· ···················· ····················
R 176 hCoV-19/England/CAMC-B2143F/2020|EPI_ISL_647454|20 ···················· ···················· ···················· ···················· ···················· ····················
R 177 hCoV-19/England/CAMC-B22D33/2020|EPI_ISL_647455|20 ···················· ···················· ···················· ···················· ···················· ····················
R 178 hCoV-19/England/CAMC-B22BDF/2020|EPI_ISL_647456|20 ···················· ···················· ···················· ···················· ···················· ····················
R 179 hCoV-19/England/CAMC-B20FE4/2020|EPI_ISL_647457|20 ···················· ···················· ···················· ···················· ···················· ····················
R 180 hCoV-19/England/CAMC-B22C45/2020|EPI_ISL_647458|20 ···················· ···················· ···················· ···················· ···················· ····················
R 181 hCoV-19/England/CAMC-B219F4/2020|EPI_ISL_647459|20 ···················· ···················· ···················· ···················· ···················· ····················
R 182 hCoV-19/England/CAMC-B21CBF/2020|EPI_ISL_647460|20 ···················· ···················· ···················· ···················· ····T··············· ····················
R 183 hCoV-19/England/CAMC-B21A88/2020|EPI_ISL_647461|20 ···················· ···················· ···················· ···················· ···················· ····················
R 184 hCoV-19/England/CAMC-B22780/2020|EPI_ISL_647462|20 ···················· ···················· ···················· ···················· ···················· ····················
R 185 hCoV-19/England/CAMC-B20FD5/2020|EPI_ISL_647463|20 ···················· ···················· ···················· ···················· ···················· ····················
R 186 hCoV-19/England/CAMC-B229B7/2020|EPI_ISL_647466|20 ···················· ·A·······A·········· ···················· ···················· ···················· ····················
R 187 hCoV-19/England/CAMC-B2194F/2020|EPI_ISL_647467|20 ···················· ···················· ···················· ···················· ···················· ····················
R 188 hCoV-19/England/ALDP-B1AA37/2020|EPI_ISL_647468|20 ···················· ···················· ···················· ···················· ···················· ····················
R 189 hCoV-19/England/CAMC-B1FCC3/2020|EPI_ISL_647470|20 ···················· ···················· ···················· ···················· ···················· ····················
R 190 hCoV-19/England/CAMC-B211A1/2020|EPI_ISL_647471|20 ···················· ···················· ···················· ···················· ···················· ····················
R 191 hCoV-19/England/CAMC-B20281/2020|EPI_ISL_647472|20 ···················· ···················· ···················· ···················· ···················· ····················
R 192 hCoV-19/England/ALDP-B1AA91/2020|EPI_ISL_647473|20 ···················· ···················· ···················· ···················· ···················· ····················
R 193 hCoV-19/England/ALDP-B1A8A6/2020|EPI_ISL_647474|20 ···················· ···················· ···················· ···················· ···················· ····················
R 194 hCoV-19/England/CAMC-B21217/2020|EPI_ISL_647476|20 ···················· ···················· ···················· ···················· ···················· ····················
R 195 hCoV-19/England/ALDP-B1A466/2020|EPI_ISL_647477|20 ···················· ···················· ···················· ···················· ···················· ····················
R 196 hCoV-19/England/CAMC-B1FC4B/2020|EPI_ISL_647478|20 ···················· ···················· ···················· ···················· ···················· ····················
R 197 hCoV-19/England/CAMC-B1FE90/2020|EPI_ISL_647479|20 ···················· ···················· ···················· ···················· ···················· ····················
R 198 hCoV-19/England/CAMC-B202BE/2020|EPI_ISL_647480|20 ···················· ···················· ···················· ···················· ···················· ····················
R 199 hCoV-19/England/CAMC-B206FE/2020|EPI_ISL_647481|20 ···················· ···················· ···················· ···················· ···················· ····················
R 200 hCoV-19/England/CAMC-B20E7E/2020|EPI_ISL_647482|20 ···················· ···················· ···················· ···················· ···················· ····················
R 201 hCoV-19/England/CAMC-B1FDDF/2020|EPI_ISL_647483|20 ···················· ·A·················· ···················· ···················· ···················· ···A················
R 202 hCoV-19/England/CAMC-B209C8/2020|EPI_ISL_647484|20 ···················· ···················· ···················· ···················· ···················· ····················
R 203 hCoV-19/England/CAMC-B20816/2020|EPI_ISL_647485|20 ···················· ···················· ···················· ···················· ···················· ····················
R 204 hCoV-19/England/CAMC-B20825/2020|EPI_ISL_647486|20 ···················· ···················· ···················· ···················· ···················· ····················
R 205 hCoV-19/England/CAMC-B20CDE/2020|EPI_ISL_647487|20 ···················· ···················· ···················· ···················· ···················· ····················
R 206 hCoV-19/England/CAMC-B1FC0F/2020|EPI_ISL_647489|20 ···················· ···················· ···················· ···················· ···················· ····················
R 207 hCoV-19/England/CAMC-B20C92/2020|EPI_ISL_647490|20 ···················· ···················· ···················· ···················· ···················· ····················
R 208 hCoV-19/England/CAMC-B21156/2020|EPI_ISL_647491|20 ···················· ···················· ···················· ···················· ···················· ····················
R 209 hCoV-19/England/CAMC-B21147/2020|EPI_ISL_647492|20 ···················· ···················· ···················· ···················· ···················· ····················
R 210 hCoV-19/England/CAMC-B21305/2020|EPI_ISL_647493|20 ···················· ···················· ···················· ···················· ···················· ····················
R 211 hCoV-19/England/CAMC-B20E14/2020|EPI_ISL_647494|20 ···················· ···················· ···················· ···················· ···················· ····················
R 212 hCoV-19/England/CAMC-B2056A/2020|EPI_ISL_647495|20 ···················· ···················· ···················· ···················· ····T··············· ····················
R 213 hCoV-19/England/CAMC-B1FE63/2020|EPI_ISL_647496|20 ···················· ···················· ···················· ···················· ······T············· ····················
R 214 hCoV-19/England/ALDP-B1A4B1/2020|EPI_ISL_647497|20 ···················· ···················· ···················· ···················· ···················· ····················
R 215 hCoV-19/England/CAMC-B20861/2020|EPI_ISL_647498|20 ···················· ···················· ···················· ···················· ···················· ····················
R 216 hCoV-19/England/ALDP-B1A4C0/2020|EPI_ISL_647499|20 ···················· ···················· ···················· ···················· ···················· ····················
R 217 hCoV-19/Singapore/1275/2020|EPI_ISL_648760|2020-05 ···················· ···················· ···················· ···················· ···················· ····················
R 218 hCoV-19/Singapore/1276/2020|EPI_ISL_648761|2020-05 ···················· ···················· ···················· ···················· ···················· ····················
R 219 hCoV-19/Singapore/1277/2020|EPI_ISL_648762|2020-05 ···················· ···················· ···················· ···················· ···················· ····················
R 220 hCoV-19/Singapore/1278/2020|EPI_ISL_648763|2020-05 ···················· ···················· ···················· ···················· ···················· ····················

R 221 hCoV-19/Singapore/1279/2020|EPI_ISL_648764|2020-05 ···················· ···················· ····················· ···················· ···················· ····················
R 222 hCoV-19/Singapore/1280/2020|EPI_ISL_648765|2020-05 ···················· ···················· ····················· ···················· ···················· ····················
R 223 hCoV-19/Singapore/1281/2020|EPI_ISL_648766|2020-05 ···················· ···················· ····················· ···················· ···················· ····················
R 224 hCoV-19/Singapore/1282/2020|EPI_ISL_648767|2020-05 ···················· ···················· ····················· ···················· ···················· ····················
R 225 hCoV-19/Singapore/1283/2020|EPI_ISL_648768|2020-05 ···················· ···················· ····················· ···················· ···················· ····················
R 226 hCoV-19/Singapore/1284/2020|EPI_ISL_648769|2020-05 ···················· ···················· ····················· ···················· ···················· ····················
R 227 hCoV-19/Singapore/1285/2020|EPI_ISL_648770|2020-05 ···················· ···················· ····················· ···················· ···················· ····················
R 228 hCoV-19/Singapore/1286/2020|EPI_ISL_648771|2020-05 ···················· ···················· ····················· ···················· ···················· ····················
R 229 hCoV-19/Singapore/1287/2020|EPI_ISL_648772|2020-05 ···················· ···················· ····················· ···················· ···················· ····················
R 230 hCoV-19/Singapore/1288/2020|EPI_ISL_648773|2020-05 ···················· ···················· ····················· ···················· ···················· ····················
R 231 hCoV-19/Singapore/1289/2020|EPI_ISL_648774|2020-05 ···················· ···················· ····················· ···················· ···················· ····················
R 232 hCoV-19/Singapore/1290/2020|EPI_ISL_648775|2020-05 ···················· ···················· ····················· ···················· ···················· ····················
R 233 hCoV-19/Singapore/1291/2020|EPI_ISL_648776|2020-05 ···················· ···················· ····················· ···················· ···················· ····················
R 234 hCoV-19/Singapore/1292/2020|EPI_ISL_648777|2020-05 ···················· ···················· ····················· ···················· ···················· ····················
R 235 hCoV-19/Singapore/1293/2020|EPI_ISL_648778|2020-05 ···················· ···················· ····················· ···················· ···················· ····················
R 236 hCoV-19/Singapore/1294/2020|EPI_ISL_648779|2020-05 ···················· ···················· ····················· ···················· ···················· ····················
R 237 hCoV-19/Singapore/1295/2020|EPI_ISL_648780|2020-05 ···················· ···················· ····················· ···················· ···················· ····················
R 238 hCoV-19/Singapore/1296/2020|EPI_ISL_648781|2020-05 ···················· ···················· ····················· ···················· ···················· ····················
R 239 hCoV-19/Singapore/1297/2020|EPI_ISL_648782|2020-05 ···················· ···················· ····················· ···················· ···················· ····················
R 240 hCoV-19/Singapore/1298/2020|EPI_ISL_648783|2020-05 ···················· ···················· ····················· ···················· ···················· ····················
R 241 hCoV-19/Singapore/1299/2020|EPI_ISL_648784|2020-05 ···················· ···················· ····················· ···················· ···················· ····················
R 242 hCoV-19/Singapore/668nan/2020|EPI_ISL_648785|2020- ···················· ···················· ····················· ···················· ···················· ····················
R 243 hCoV-19/Singapore/1410/2020|EPI_ISL_648786|2020-05 ···················· ···················· ····················· ···················· ···················· ····················
R 244 hCoV-19/Singapore/1300/2020|EPI_ISL_648787|2020-05 ···················· ···················· ····················· ···················· ···················· ····················
R 245 hCoV-19/Singapore/1301/2020|EPI_ISL_648788|2020-05 ···················· ···················· ····················· ···················· ···················· ····················
R 246 hCoV-19/Singapore/1302/2020|EPI_ISL_648789|2020-05 ···················· ···················· ····················· ···················· ···················· ····················
R 247 hCoV-19/Singapore/1303/2020|EPI_ISL_648790|2020-05 ···················· ···················· ····················· ···················· ···················· ····················
R 248 hCoV-19/Singapore/1304/2020|EPI_ISL_648791|2020-05 ···················· ···················· ····················· ···················· ···················· ····················
R 249 hCoV-19/Singapore/1305/2020|EPI_ISL_648792|2020-05 ···················· ···················· ····················· ···················· ···················· ····················
R 250 hCoV-19/Singapore/1306/2020|EPI_ISL_648793|2020-05 ···················· ···················· ····················· ···················· ···················· ····················
R 251 hCoV-19/Singapore/1307/2020|EPI_ISL_648794|2020-05 ···················· ···················· ····················· ···················· ···················· ····················
R 252 hCoV-19/Singapore/1308/2020|EPI_ISL_648795|2020-05 ···················· ···················· ····················· ···················· ···················· ····················
R 253 hCoV-19/Singapore/1309/2020|EPI_ISL_648796|2020-05 ···················· ···················· ····················· ···················· ···················· ····················
R 254 hCoV-19/Singapore/1310/2020|EPI_ISL_648797|2020-05 ···················· ···················· ····················· ···················· ···················· ····················
R 255 hCoV-19/Singapore/1311/2020|EPI_ISL_648798|2020-05 ···················· ···················· ····················· ···················· ···················· ····················
R 256 hCoV-19/Singapore/1312/2020|EPI_ISL_648799|2020-05 ···················· ···················· ····················· ···················· ···················· ····················
R 258 hCoV-19/Australia/VIC12073/2020|EPI_ISL_641209|202 ···················· ···················· ····················· ···················· ···················· ····················
R 259 hCoV-19/Australia/VIC15969/2020|EPI_ISL_641211|202 ···················· ···················· ····················· ···················· ···················· ····················
R 260 hCoV-19/Australia/VIC16487/2020|EPI_ISL_641212|202 ···················· ···················· ····················· ···················· ···················· ····················
R 262 hCoV-19/Australia/VIC11780/2020|EPI_ISL_641216|202 ···················· ···················· ····················· ···················· ···················· ····················
R 264 hCoV-19/Australia/VIC12669/2020|EPI_ISL_641219|202 ···················· ···················· ····················· ···················· ···················· ····················
R 265 hCoV-19/Australia/VIC16180/2020|EPI_ISL_641221|202 ···················· ···················· ····················· ···················· ···················· ····················
R 266 hCoV-19/Australia/VIC16343/2020|EPI_ISL_641222|202 ···················· ···················· ····················· ···················· ···················· ····················
R 267 hCoV-19/Australia/VIC16437/2020|EPI_ISL_641223|202 ···················· ···················· ····················· ···················· ···················· ····················
R 268 hCoV-19/Australia/VIC16529/2020|EPI_ISL_641224|202 ···················· ···················· ····················· ···················· ···················· ····················
R 269 hCoV-19/Australia/VIC12355/2020|EPI_ISL_641225|202 ···················· ···················· ····················· ···················· ···················· ····················
R 270 hCoV-19/Australia/VIC12657/2020|EPI_ISL_641226|202 ···················· ···················· ····················· ···················· ···················· ····················
R 271 hCoV-19/Australia/VIC12117/2020|EPI_ISL_641228|202 ···················· ···················· ····················· ···················· ···················· ····················
R 272 hCoV-19/Australia/VIC12788/2020|EPI_ISL_641229|202 ···················· ······A············· ····················· ···················· ···················· ····················
R 274 hCoV-19/Australia/VIC12046/2020|EPI_ISL_641233|202 ···················· ···················· ····················· ···················· ···················· ····················
R 275 hCoV-19/Australia/VIC16396/2020|EPI_ISL_641234|202 ···················· ···················· ····················· ···················· ···················· ····················
R 276 hCoV-19/Australia/VIC12753/2020|EPI_ISL_641235|202 ···················· ···················· ····················· ···················· ···················· ····················
R 277 hCoV-19/Australia/VIC12687/2020|EPI_ISL_641238|202 ···················· ···················· ····················· ···················· ···················· ····················
R 278 hCoV-19/Australia/VIC12754/2020|EPI_ISL_641239|202 ···················· ···················· ····················· ···················· ···················· ····················
R 279 hCoV-19/Australia/VIC12775/2020|EPI_ISL_641240|202 ···················· ···················· ····················· ···················· ···················· ····················
R 280 hCoV-19/Australia/VIC16379/2020|EPI_ISL_641241|202 ···················· ···················· ····················· ···················· ···················· ····················
R 281 hCoV-19/Australia/VIC12075/2020|EPI_ISL_641242|202 ···················· ···················· ····················· ···················· ···················· ····················
R 284 hCoV-19/Australia/VIC12862/2020|EPI_ISL_641245|202 ···················· ···················· ····················· ···················· ···················· ····················
R 285 hCoV-19/Australia/VIC12180/2020|EPI_ISL_641248|202 ···················· ···················· ····················· ···················· ···················· ····················
R 286 hCoV-19/Australia/VIC16539/2020|EPI_ISL_641249|202 ···················· ···················· ····················· ···················· ···················· ····················
R 287 hCoV-19/Australia/VIC12421/2020|EPI_ISL_641250|202 ···················· ···················· ····················· ···················· ···················· ····················
R 288 hCoV-19/Australia/VIC12878/2020|EPI_ISL_641252|202 ···················· ···················· ····················· ···················· ···················· ····················
R 290 hCoV-19/Australia/VIC16357/2020|EPI_ISL_641254|202 ···················· ···················· ····················· ···················· ···················· ····················
R 291 hCoV-19/Australia/VIC16065/2020|EPI_ISL_641255|202 ···················· ···················· ····················· ···················· ···················· ····················
R 293 hCoV-19/Australia/VIC16309/2020|EPI_ISL_641257|202 ···················· ···················· ····················· ···················· ···················· ····················
R 294 hCoV-19/Australia/VIC16519/2020|EPI_ISL_641259|202 ···················· ···················· ····················· ···················· ···················· NN··················
R 295 hCoV-19/Australia/VIC16123/2020|EPI_ISL_641261|202 ···················· ···················· ····················· ···················· ···················· ····················
R 296 hCoV-19/Australia/VIC12459/2020|EPI_ISL_641262|202 ···················· ···················· ····················· ···················· ···················· ····················
R 297 hCoV-19/Australia/VIC15932/2020|EPI_ISL_641263|202 ···················· ···················· ····················· ···················· ···················· ····················
R 298 hCoV-19/Australia/VIC11626/2020|EPI_ISL_641264|202 ···················· ···················· ····················· ···················· ···················· ····················
R 300 hCoV-19/Australia/VIC12717/2020|EPI_ISL_641267|202 ···················· ······A············· ····················· ···················· ···················· ····················
R 301 hCoV-19/Australia/VIC16568/2020|EPI_ISL_641268|202 ···················· ···················· ····················· ···················· ···················· ····················
R 302 hCoV-19/Australia/VIC12423/2020|EPI_ISL_641270|202 ···················· ···················· ····················· ···················· ···················· ····················
R 303 hCoV-19/Australia/VIC13265/2020|EPI_ISL_641271|202 ···················· ···················· ····················· ···················· ···················· ····················
R 304 hCoV-19/Australia/VIC12592/2020|EPI_ISL_641272|202 ···················· ···················· ····················· ···················· ···················· ····················
R 305 hCoV-19/Australia/VIC12556/2020|EPI_ISL_641274|202 ···················· ···················· ····················· ···················· ···················· ····················
R 306 hCoV-19/Australia/VIC16572/2020|EPI_ISL_641275|202 ···················· ···················· ····················· ···················· ···················· ····················
R 309 hCoV-19/Australia/VIC13254/2020|EPI_ISL_641281|202 ···················· ···················· ····················· ···················· ···················· ····················
R 310 hCoV-19/Australia/VIC12467/2020|EPI_ISL_641282|202 ···················· ···················· ····················· ···················· ···················· ····················
R 311 hCoV-19/Australia/VIC12852/2020|EPI_ISL_641283|202 ···················· ···················· ····················· ···················· ···················· ····················

| | | | | | | | |
|---|---|---|---|---|---|---|---|
| R | 312 hCoV-19/Australia/VIC15912/2020\|EPI_ISL_641284\|202 | ···················· | ···················· | ···················· | ···················· | ···················· | ···················· |
| R | 313 hCoV-19/Australia/VIC16017/2020\|EPI_ISL_641285\|202 | ···················· | ···················· | ···················· | ···················· | ···················· | ···················· |
| R | 314 hCoV-19/Australia/VIC13075/2020\|EPI_ISL_641286\|202 | ···················· | ···················· | ···················· | ···················· | ···················· | ···················· |
| R | 315 hCoV-19/Australia/VIC16083/2020\|EPI_ISL_641287\|202 | ···················· | ···················· | ···················· | ···················· | ···················· | ···················· |
| R | 316 hCoV-19/Australia/VIC12154/2020\|EPI_ISL_641289\|202 | ···················· | ···················· | ···················· | ···················· | ···················· | ···················· |
| R | 317 hCoV-19/Australia/VIC15928/2020\|EPI_ISL_641300\|202 | ···················· | ···················· | ···················· | ···················· | ···················· | ···················· |
| R | 318 hCoV-19/Australia/VIC16038/2020\|EPI_ISL_641301\|202 | ···················· | ···················· | ···················· | ···················· | ···················· | ···················· |
| R | 319 hCoV-19/England/QEUH-B1443F/2020\|EPI_ISL_642534\|20 | ···················· | ···················· | ···················· | ···················· | ···················· | ···················· |
| R | 320 hCoV-19/England/QEUH-B11DA0/2020\|EPI_ISL_642535\|20 | ···················· | ···················· | ···················· | ···················· | ···················· | ···················· |
| R | 321 hCoV-19/England/QEUH-B14402/2020\|EPI_ISL_642536\|20 | ···················· | ···················· | ···················· | ···················· | ···················· | ···················· |
| R | 322 hCoV-19/England/QEUH-B125A7/2020\|EPI_ISL_642537\|20 | ···················· | ···················· | ···················· | ···················· | ···················· | ···················· |
| R | 323 hCoV-19/England/QEUH-B11265/2020\|EPI_ISL_642538\|20 | ···················· | ···················· | ···················· | ···················· | ···················· | ···················· |
| R | 324 hCoV-19/England/QEUH-B0F21F/2020\|EPI_ISL_642539\|20 | ···················· | ···················· | ···················· | ···················· | ···················· | ···················· |
| R | 325 hCoV-19/England/QEUH-B12B5A/2020\|EPI_ISL_642540\|20 | ···················· | ···················· | ···················· | ···················· | ···················· | ···················· |
| R | 326 hCoV-19/England/QEUH-B11E25/2020\|EPI_ISL_642541\|20 | ···················· | ···················· | ···················· | ···················· | ······T············· | ···················· |
| R | 327 hCoV-19/England/QEUH-B0F18B/2020\|EPI_ISL_642542\|20 | ···················· | ···················· | ···················· | ···················· | ···················· | ···················· |
| R | 328 hCoV-19/England/QEUH-B1006C/2020\|EPI_ISL_642545\|20 | ···················· | ···················· | ···················· | ···················· | ···················· | ···················· |
| R | 329 hCoV-19/England/QEUH-B1004E/2020\|EPI_ISL_642546\|20 | ···················· | ···················· | ···················· | ···················· | ···················· | ···················· |
| R | 330 hCoV-19/England/QEUH-B12668/2020\|EPI_ISL_642547\|20 | ···················· | ···················· | ···················· | ···················· | ···················· | ···················· |
| R | 331 hCoV-19/England/QEUH-B1204C/2020\|EPI_ISL_642548\|20 | ···················· | ···················· | ···················· | ···················· | ···················· | ···················· |
| R | 332 hCoV-19/England/QEUH-B1238F/2020\|EPI_ISL_642550\|20 | ···················· | ···················· | ···················· | ···················· | ···················· | ···················· |
| R | 333 hCoV-19/England/QEUH-B109E9/2020\|EPI_ISL_642551\|20 | ···················· | ···················· | ···················· | ···················· | ···················· | ···················· |
| R | 334 hCoV-19/England/QEUH-B14420/2020\|EPI_ISL_642552\|20 | ···················· | ···················· | ···················· | ···················· | ···················· | ···················· |
| R | 335 hCoV-19/England/QEUH-B1099E/2020\|EPI_ISL_642553\|20 | ···················· | ···················· | ···················· | ···················· | ···················· | ···················· |
| R | 336 hCoV-19/England/QEUH-B124D7/2020\|EPI_ISL_642554\|20 | ···················· | ···················· | ···················· | ···················· | ···················· | ···················· |
| R | 337 hCoV-19/England/QEUH-B1007B/2020\|EPI_ISL_642555\|20 | ·····G·············· | ···················· | ···················· | ···················· | ···················· | ···················· |
| R | 338 hCoV-19/England/QEUH-B125F2/2020\|EPI_ISL_642556\|20 | ···················· | ···················· | ···················· | ···················· | ···················· | ···················· |
| R | 339 hCoV-19/England/QEUH-B100B7/2020\|EPI_ISL_642557\|20 | ···················· | ···················· | ···················· | ···················· | ···················· | ···················· |
| R | 340 hCoV-19/England/QEUH-B108DD/2020\|EPI_ISL_642558\|20 | ···················· | ···················· | ···················· | ···················· | ···················· | ···················· |
| R | 341 hCoV-19/England/QEUH-B10952/2020\|EPI_ISL_642559\|20 | ···················· | ···················· | ···················· | ···················· | ···················· | ···················· |
| R | 342 hCoV-19/England/QEUH-B11362/2020\|EPI_ISL_642560\|20 | ···················· | ···················· | ···················· | ···················· | ···················· | ···················· |
| R | 343 hCoV-19/England/QEUH-B0F1A9/2020\|EPI_ISL_642561\|20 | ···················· | ···················· | ···················· | ···················· | ···················· | ···················· |
| R | 344 hCoV-19/England/QEUH-B1254D/2020\|EPI_ISL_642562\|20 | ···················· | ···················· | ···················· | ···················· | ···················· | ···················· |
| R | 345 hCoV-19/England/QEUH-B11274/2020\|EPI_ISL_642563\|20 | ···················· | ···················· | ···················· | ···················· | ······T············· | ···················· |
| R | 346 hCoV-19/England/QEUH-B12B69/2020\|EPI_ISL_642564\|20 | ···················· | ···················· | ···················· | ···················· | ···················· | ···················· |
| R | 347 hCoV-19/England/QEUH-B12A7B/2020\|EPI_ISL_642565\|20 | ···················· | ···················· | ···················· | ···················· | ···················· | ···················· |
| R | 348 hCoV-19/England/QEUH-B12DFA/2020\|EPI_ISL_642566\|20 | ···················· | ···················· | ···················· | ···················· | ···················· | ···················· |
| R | 349 hCoV-19/England/QEUH-B10970/2020\|EPI_ISL_642567\|20 | ···················· | ···················· | ···················· | ···················· | ···················· | ···················· |
| R | 350 hCoV-19/England/QEUH-B143C9/2020\|EPI_ISL_642568\|20 | ···················· | ···················· | ···················· | ···················· | ···················· | ···················· |
| R | 351 hCoV-19/England/QEUH-B12B78/2020\|EPI_ISL_642569\|20 | ···················· | ···················· | ···················· | ···················· | ···················· | ···················· |
| R | 352 hCoV-19/England/QEUH-B11317/2020\|EPI_ISL_642570\|20 | ···················· | ···················· | ···················· | ···················· | ···················· | ···················· |
| R | 353 hCoV-19/England/QEUH-B10B98/2020\|EPI_ISL_642571\|20 | ···················· | ······A············· | ···················· | ···················· | ···················· | ···················· |
| R | 354 hCoV-19/England/QEUH-B12B3C/2020\|EPI_ISL_642572\|20 | ···················· | ·A·················· | ···················· | ···················· | ···················· | ···A················ |
| R | 355 hCoV-19/England/QEUH-B12E15/2020\|EPI_ISL_642574\|20 | ···················· | ···················· | ···················· | ···················· | ···················· | ···················· |
| R | 356 hCoV-19/England/QEUH-B144C6/2020\|EPI_ISL_642575\|20 | ···················· | ···················· | ···················· | ···················· | ···················· | ···················· |
| R | 357 hCoV-19/England/QEUH-B11283/2020\|EPI_ISL_642576\|20 | ···················· | ···················· | ···················· | ···················· | ···················· | ···················· |
| R | 358 hCoV-19/England/QEUH-B1010F/2020\|EPI_ISL_642578\|20 | ···················· | ···················· | ···················· | ···················· | ···················· | ···················· |
| R | 359 hCoV-19/England/QEUH-B1445D/2020\|EPI_ISL_642579\|20 | ···················· | ···················· | ···················· | ···················· | ······T············· | ···················· |
| R | 360 hCoV-19/England/QEUH-B11238/2020\|EPI_ISL_642580\|20 | ···················· | ···················· | ···················· | ···················· | ···················· | ···················· |
| R | 361 hCoV-19/England/QEUH-B129E7/2020\|EPI_ISL_642581\|20 | ···················· | ···················· | ···················· | ···················· | ···················· | ···················· |
| R | 362 hCoV-19/England/QEUH-B10BD4/2020\|EPI_ISL_642584\|20 | ···················· | ···················· | ···················· | ···················· | ···················· | ···················· |
| R | 363 hCoV-19/England/QEUH-B10B2F/2020\|EPI_ISL_642585\|20 | ···················· | ······A············· | ···················· | ···················· | ···················· | ···················· |
| R | 364 hCoV-19/England/QEUH-B0ED1F/2020\|EPI_ISL_642586\|20 | ···················· | ···················· | ···················· | ···················· | ···················· | ···················· |
| R | 365 hCoV-19/England/QEUH-B0F24C/2020\|EPI_ISL_642587\|20 | ···················· | ···················· | ···················· | ···················· | ···················· | ···················· |
| R | 366 hCoV-19/England/QEUH-B10BC5/2020\|EPI_ISL_642588\|20 | ···················· | ···················· | ···················· | ···················· | ···················· | ···················· |
| R | 367 hCoV-19/England/QEUH-B11353/2020\|EPI_ISL_642589\|20 | ···················· | ···················· | ···················· | ···················· | ···················· | ···················· |
| R | 368 hCoV-19/England/QEUH-B144E4/2020\|EPI_ISL_642590\|20 | ···················· | ···················· | ···················· | ···················· | ···················· | ···················· |
| R | 369 hCoV-19/England/QEUH-B12C2A/2020\|EPI_ISL_642591\|20 | ···················· | ···················· | ···················· | ···················· | ···················· | ···················· |
| R | 370 hCoV-19/England/QEUH-B109F8/2020\|EPI_ISL_642593\|20 | ···················· | ·A·················· | ···················· | ···················· | ···················· | ···················· |
| R | 371 hCoV-19/England/QEUH-B12B2D/2020\|EPI_ISL_642594\|20 | ···················· | ···················· | ···················· | ···················· | ···················· | ···················· |
| R | 372 hCoV-19/England/QEUH-B0FFFA/2020\|EPI_ISL_642595\|20 | ···················· | ···················· | ···················· | ···················· | ···················· | ···················· |
| R | 373 hCoV-19/England/QEUH-B12079/2020\|EPI_ISL_642596\|20 | ···················· | ···················· | ···················· | ···················· | ···················· | ···················· |
| R | 374 hCoV-19/England/QEUH-B12431/2020\|EPI_ISL_642597\|20 | ···················· | ···················· | ···················· | ···················· | ···················· | ···················· |
| R | 375 hCoV-19/England/QEUH-B12817/2020\|EPI_ISL_642598\|20 | ···················· | ···················· | ···················· | ···················· | ···················· | ···················· |
| R | 376 hCoV-19/England/QEUH-B12B4B/2020\|EPI_ISL_642599\|20 | ···················· | ···················· | ···················· | ···················· | ···················· | ···················· |
| R | 377 hCoV-19/England/QEUH-B112ED/2020\|EPI_ISL_642600\|20 | ···················· | ···················· | ···················· | ···················· | ···················· | ···A················ |
| R | 378 hCoV-19/England/QEUH-B113AE/2020\|EPI_ISL_642601\|20 | ···················· | ···················· | ···················· | ···················· | ···················· | ···················· |
| R | 379 hCoV-19/England/QEUH-B12A4E/2020\|EPI_ISL_642603\|20 | ···················· | ···················· | ···················· | ···················· | ······T············· | ···················· |
| R | 380 hCoV-19/England/QEUH-B11247/2020\|EPI_ISL_642604\|20 | ···················· | ···················· | ···················· | ···················· | ···················· | ···················· |
| R | 381 hCoV-19/England/QEUH-B11FB9/2020\|EPI_ISL_642605\|20 | ···················· | ···················· | ···················· | ···················· | ···················· | ···················· |
| R | 382 hCoV-19/England/QEUH-B12088/2020\|EPI_ISL_642608\|20 | ···················· | ···················· | ···················· | ···················· | ···················· | ···················· |
| R | 383 hCoV-19/England/QEUH-B124C8/2020\|EPI_ISL_642609\|20 | ···················· | ···················· | ···················· | ···················· | ···················· | ···················· |
| R | 384 hCoV-19/England/QEUH-B11DCE/2020\|EPI_ISL_642610\|20 | ···················· | ···················· | ···················· | ···················· | ···················· | ···················· |
| R | 385 hCoV-19/England/QEUH-B10B3E/2020\|EPI_ISL_642624\|20 | ···················· | ···················· | ···················· | ···················· | ···················· | ···················· |
| R | 386 hCoV-19/England/QEUH-B143D8/2020\|EPI_ISL_642630\|20 | ···················· | ···················· | ···················· | ···················· | ····T··············· | ···················· |
| R | 387 hCoV-19/England/QEUH-B0EE2B/2020\|EPI_ISL_642631\|20 | ···················· | ···················· | ···················· | ···················· | ···················· | ···················· |
| R | 388 hCoV-19/England/QEUH-B10B7A/2020\|EPI_ISL_642632\|20 | ···················· | ···················· | ···················· | ···················· | ···················· | ···················· |
| R | 389 hCoV-19/England/CAMC-B083D7/2020\|EPI_ISL_643865\|20 | ···················· | ···················· | ···················· | ···················· | ······T············· | ···················· |
| R | 390 hCoV-19/England/CAMC-B071DE/2020\|EPI_ISL_643866\|20 | ···················· | ···················· | ···················· | ···················· | ···················· | ···················· |
| R | 391 hCoV-19/England/CAMC-B0894E/2020\|EPI_ISL_643867\|20 | ···················· | ···················· | ···················· | ···················· | ···················· | ···················· |

| | | | | | | | |
|---|---|---|---|---|---|---|---|
| R | 392 hCoV-19/England/CAMC-B074A8/2020\|EPI_ISL_643868\|20 | ···················· | ···················· | ···················· | ···················· | ···················· | ···················· |
| R | 393 hCoV-19/England/CAMC-B082AD/2020\|EPI_ISL_643869\|20 | ···················· | ···················· | ···················· | ···················· | ···················· | ···················· |
| R | 394 hCoV-19/England/CAMC-B07350/2020\|EPI_ISL_643870\|20 | ···················· | ···················· | ···················· | ···················· | ···················· | ···················· |
| R | 395 hCoV-19/England/CAMC-B07A5B/2020\|EPI_ISL_643871\|20 | ···················· | ···················· | ···················· | ···················· | ·····T·············· | ···················· |
| R | 396 hCoV-19/England/CAMC-B08270/2020\|EPI_ISL_643872\|20 | ···················· | ···················· | ···················· | ···················· | ···················· | ···················· |
| R | 397 hCoV-19/England/CAMC-B07CA0/2020\|EPI_ISL_643873\|20 | ···················· | ···················· | ···················· | ···················· | ···················· | ···················· |
| R | 398 hCoV-19/England/CAMC-B07C91/2020\|EPI_ISL_643874\|20 | ···················· | ···················· | ···················· | ···················· | ···················· | ···················· |
| R | 399 hCoV-19/England/CAMC-B07A6A/2020\|EPI_ISL_643875\|20 | ···················· | ···················· | ···················· | ···················· | ···················· | ···················· |
| R | 400 hCoV-19/England/CAMC-B078F7/2020\|EPI_ISL_643876\|20 | ···················· | ···················· | ···················· | ···················· | ·····T·············· | ···················· |
| R | 401 hCoV-19/England/CAMC-B08850/2020\|EPI_ISL_643878\|20 | ···················· | ···················· | ···················· | ···················· | ···················· | ···················· |
| R | 402 hCoV-19/England/CAMC-B07183/2020\|EPI_ISL_643879\|20 | ···················· | ···················· | ···················· | ···················· | ···················· | ···················· |
| R | 403 hCoV-19/England/CAMC-B083AA/2020\|EPI_ISL_643880\|20 | ···················· | ···················· | ···················· | ···················· | ···················· | ···················· |
| R | 404 hCoV-19/England/CAMC-B07F01/2020\|EPI_ISL_643881\|20 | ···················· | ···················· | ···················· | ···················· | ···················· | ···················· |
| R | 405 hCoV-19/England/CAMC-B074B7/2020\|EPI_ISL_643882\|20 | ···················· | ···················· | ···················· | ···················· | ···················· | ···················· |
| R | 406 hCoV-19/England/CAMC-B07833/2020\|EPI_ISL_643883\|20 | ···················· | ···················· | ···················· | ···················· | ···················· | ···················· |
| R | 407 hCoV-19/England/CAMC-B075F0/2020\|EPI_ISL_643885\|20 | ···················· | ···················· | ···················· | ···················· | ···················· | ···················· |
| R | 408 hCoV-19/England/CAMC-B08CEB/2020\|EPI_ISL_643886\|20 | ···················· | ···················· | ···················· | ···················· | ···················· | ···················· |
| R | 409 hCoV-19/England/CAMC-B077BE/2020\|EPI_ISL_643887\|20 | ···················· | ···················· | ···················· | ···················· | ·····T·············· | ···················· |
| R | 410 hCoV-19/England/CAMC-B073AB/2020\|EPI_ISL_643888\|20 | ···················· | ···················· | ···················· | ···················· | ···················· | ···················· |
| R | 411 hCoV-19/England/CAMC-B08304/2020\|EPI_ISL_643889\|20 | ···················· | ···················· | ···················· | ···················· | ···················· | ···················· |
| R | 412 hCoV-19/England/CAMC-B07156/2020\|EPI_ISL_643890\|20 | ···················· | ···················· | ···················· | ···················· | ···················· | ···················· |
| R | 413 hCoV-19/England/CAMC-B089D5/2020\|EPI_ISL_643893\|20 | ···················· | ···················· | ···················· | ···················· | ···················· | ···················· |
| R | 414 hCoV-19/England/CAMC-B07718/2020\|EPI_ISL_643894\|20 | ···················· | ···················· | ···················· | ···················· | ·····T·············· | ···················· |
| R | 415 hCoV-19/England/CAMC-B07C73/2020\|EPI_ISL_643895\|20 | ···················· | ···················· | ···················· | ···················· | ···················· | ···················· |
| R | 416 hCoV-19/England/CAMC-B089F3/2020\|EPI_ISL_643896\|20 | ···················· | ···················· | ···················· | ···················· | ···················· | ···················· |
| R | 417 hCoV-19/England/CAMC-B0799A/2020\|EPI_ISL_643897\|20 | ···················· | ···················· | ···················· | ···················· | ···················· | ···················· |
| R | 418 hCoV-19/England/CAMC-B07CCE/2020\|EPI_ISL_643899\|20 | ···················· | ···················· | ···················· | ···················· | ·····T·············· | ···················· |
| R | 419 hCoV-19/England/CAMC-B087DB/2020\|EPI_ISL_643901\|20 | ···················· | ···················· | ···················· | ···················· | ···················· | ···················· |
| R | 420 hCoV-19/England/CAMC-B0837D/2020\|EPI_ISL_643903\|20 | ···················· | ···················· | ···················· | ···················· | ···················· | ···················· |
| R | 421 hCoV-19/England/CAMC-B07217/2020\|EPI_ISL_643904\|20 | ···················· | ···················· | ···················· | ···················· | ·····T·············· | ···················· |
| R | 422 hCoV-19/England/CAMC-B081BF/2020\|EPI_ISL_643905\|20 | ···················· | ···················· | ···················· | ···················· | ···················· | ···················· |
| R | 423 hCoV-19/England/CAMC-B08920/2020\|EPI_ISL_643906\|20 | ···················· | ···················· | ···················· | ···················· | ···················· | ···················· |
| R | 424 hCoV-19/England/CAMC-B07262/2020\|EPI_ISL_643908\|20 | ···················· | ·A·················· | ···················· | ···················· | ···················· | ··A················· |
| R | 425 hCoV-19/England/CAMC-B0755A/2020\|EPI_ISL_643909\|20 | ···················· | ·A·················· | ···················· | ···················· | ···················· | ···················· |
| R | 426 hCoV-19/England/CAMC-B07314/2020\|EPI_ISL_643910\|20 | ···················· | ···················· | ···················· | ···················· | ···················· | ···················· |
| R | 427 hCoV-19/England/CAMC-B0861A/2020\|EPI_ISL_643911\|20 | ···················· | ···················· | ···················· | ···················· | ···················· | ···················· |
| R | 428 hCoV-19/England/CAMC-B08137/2020\|EPI_ISL_643912\|20 | ···················· | ···················· | ···················· | ···················· | ···················· | ···················· |
| R | 429 hCoV-19/England/CAMC-B073C9/2020\|EPI_ISL_643913\|20 | ···················· | ···················· | ···················· | ···················· | ·····T·············· | ···················· |
| R | 430 hCoV-19/England/CAMC-B08C09/2020\|EPI_ISL_643914\|20 | ···················· | ···················· | ···················· | ···················· | ···················· | ···················· |
| R | 431 hCoV-19/England/CAMC-B0744E/2020\|EPI_ISL_643916\|20 | ···················· | ···················· | ···················· | ···················· | ···················· | ···················· |
| R | 432 hCoV-19/England/CAMC-B07B67/2020\|EPI_ISL_643917\|20 | ···················· | ···················· | ···················· | ···················· | ···················· | ···················· |
| R | 433 hCoV-19/England/CAMC-B073D8/2020\|EPI_ISL_643918\|20 | ···················· | ···················· | ···················· | ···················· | ·····T·············· | ···················· |
| R | 434 hCoV-19/England/CAMC-B08AB4/2020\|EPI_ISL_643919\|20 | ···················· | ···················· | ···················· | ···················· | ···················· | ···················· |
| R | 435 hCoV-19/England/CAMC-B07FC5/2020\|EPI_ISL_643920\|20 | ···················· | ···················· | ···················· | ···················· | ···················· | ···················· |
| R | 436 hCoV-19/England/CAMC-B07781/2020\|EPI_ISL_643921\|20 | ···················· | ···················· | ···················· | ···················· | ···················· | ···················· |
| R | 437 hCoV-19/England/CAMC-B07FD4/2020\|EPI_ISL_643922\|20 | ···················· | ···················· | ···················· | ···················· | ···················· | ···················· |
| R | 438 hCoV-19/England/CAMC-B088BA/2020\|EPI_ISL_643923\|20 | ···················· | ···················· | ···················· | ···················· | ···················· | ···················· |
| R | 439 hCoV-19/England/CAMC-B08A69/2020\|EPI_ISL_643924\|20 | ···················· | ···················· | ···················· | ···················· | ·····T·············· | ···················· |
| R | 440 hCoV-19/England/CAMC-B07693/2020\|EPI_ISL_643926\|20 | ···················· | ···················· | ···················· | ···················· | ···················· | ···················· |
| R | 441 hCoV-19/England/CAMC-B07D43/2020\|EPI_ISL_643927\|20 | ···················· | ···················· | ···················· | ···················· | ···················· | ···················· |
| R | 442 hCoV-19/England/CAMC-B07A79/2020\|EPI_ISL_643928\|20 | ···················· | ···················· | ···················· | ···················· | ···················· | ···················· |
| R | 443 hCoV-19/England/CAMC-B092BB/2020\|EPI_ISL_643929\|20 | ···················· | ···················· | ···················· | ···················· | ···················· | ···················· |
| R | 444 hCoV-19/England/CAMC-B082CB/2020\|EPI_ISL_643930\|20 | ···················· | ···················· | ···················· | ···················· | ···················· | ···················· |
| R | 445 hCoV-19/England/CAMC-B07253/2020\|EPI_ISL_643932\|20 | ···················· | ···················· | ···················· | ···················· | ···················· | ···················· |
| R | 446 hCoV-19/England/CAMC-B07D34/2020\|EPI_ISL_643933\|20 | ···················· | ···················· | ···················· | ···················· | ···················· | ···················· |
| R | 447 hCoV-19/England/CAMC-B088E7/2020\|EPI_ISL_643934\|20 | ···················· | ···················· | ···················· | ···················· | ···················· | ···················· |
| R | 448 hCoV-19/England/CAMC-B079E5/2020\|EPI_ISL_643935\|20 | ···················· | ···················· | ···················· | ···················· | ···················· | ···················· |
| R | 449 hCoV-19/England/CAMC-B078D9/2020\|EPI_ISL_643936\|20 | ···················· | ···················· | ···················· | ···················· | ···················· | ···················· |
| R | 450 hCoV-19/England/CAMC-B086A1/2020\|EPI_ISL_643937\|20 | ···················· | ···················· | ···················· | ···················· | ···················· | ···················· |
| R | 451 hCoV-19/England/CAMC-B07341/2020\|EPI_ISL_643938\|20 | ···················· | ···················· | ···················· | ···················· | ···················· | ···················· |
| R | 452 hCoV-19/England/CAMC-B08744/2020\|EPI_ISL_643940\|20 | ···················· | ···················· | ···················· | ···················· | ···················· | ···················· |
| R | 453 hCoV-19/England/CAMC-B07815/2020\|EPI_ISL_643941\|20 | ···················· | ···················· | ···················· | ···················· | ·····T·············· | ···················· |
| R | 454 hCoV-19/England/CAMC-B08CBE/2020\|EPI_ISL_643955\|20 | ···················· | ···················· | ···················· | ···················· | ···················· | ···················· |
| R | 455 hCoV-19/England/CAMC-B07BFE/2020\|EPI_ISL_643960\|20 | ···················· | ···················· | ···················· | ···················· | ···················· | ···················· |
| R | 456 hCoV-19/England/CAMC-B07709/2020\|EPI_ISL_643961\|20 | ···················· | ···················· | ···················· | ···················· | ···················· | ···················· |
| R | 457 hCoV-19/England/CAMC-B08340/2020\|EPI_ISL_643962\|20 | ···················· | ···················· | ···················· | ···················· | ···················· | ···················· |
| R | 458 hCoV-19/England/CAMC-B079D6/2020\|EPI_ISL_643963\|20 | ···················· | ···················· | ···················· | ···················· | ···················· | ···················· |
| R | 459 hCoV-19/England/ALDP-B23F4B/2020\|EPI_ISL_646180\|20 | ···················· | ·A·················· | ···················· | ···················· | ···················· | ···················· |
| R | 460 hCoV-19/England/ALDP-B23DD8/2020\|EPI_ISL_646182\|20 | ···················· | ···················· | ···················· | ···················· | ···················· | ···················· |
| R | 461 hCoV-19/England/QEUH-B31D7D/2020\|EPI_ISL_646183\|20 | ···················· | ···················· | ···················· | ···················· | ···················· | ···················· |
| R | 462 hCoV-19/England/QEUH-B3196A/2020\|EPI_ISL_646184\|20 | ···················· | ···················· | ···················· | ···················· | ···················· | ···················· |
| R | 463 hCoV-19/England/ALDP-B24056/2020\|EPI_ISL_646185\|20 | ···················· | ···················· | ···················· | ···················· | ···················· | ···················· |
| R | 464 hCoV-19/England/QEUH-B2DE17/2020\|EPI_ISL_646186\|20 | ···················· | ···················· | ···················· | ···················· | ···················· | ···················· |
| R | 465 hCoV-19/England/QEUH-B31ED4/2020\|EPI_ISL_646187\|20 | ···················· | ···················· | ···················· | ···················· | ···················· | ···················· |
| R | 466 hCoV-19/England/QEUH-B2E4F6/2020\|EPI_ISL_646188\|20 | ···················· | ···················· | ···················· | ···················· | ···················· | ···················· |
| R | 467 hCoV-19/England/ALDP-B23E4E/2020\|EPI_ISL_646189\|20 | ···················· | ···················· | ···················· | ···················· | ···················· | ···················· |
| R | 468 hCoV-19/England/ALDP-B23DAB/2020\|EPI_ISL_646191\|20 | ···················· | ···················· | ···················· | ···················· | ···················· | ···················· |
| R | 469 hCoV-19/England/QEUH-B2E06B/2020\|EPI_ISL_646192\|20 | ···················· | ···················· | ···················· | ···················· | ···················· | ···················· |
| R | 470 hCoV-19/England/ALDP-B23EA8/2020\|EPI_ISL_646193\|20 | ···················· | ···················· | ···················· | ···················· | ···················· | ···················· |
| R | 471 hCoV-19/England/QEUH-B31C16/2020\|EPI_ISL_646194\|20 | ···················· | ···················· | ···················· | ···················· | ···················· | ···················· |

| | | | | | | | |
|---|---|---|---|---|---|---|---|
| R | 472 hCoV-19/England/QEUH-B31AD0/2020\|EPI_ISL_646195\|20 | ···················· | ···················· | ···················· | ···················· | ···················· | ···················· |
| R | 473 hCoV-19/England/QEUH-B31979/2020\|EPI_ISL_646196\|20 | ···················· | ·A·················· | ···················· | ···················· | ···················· | ···················· |
| R | 474 hCoV-19/England/QEUH-B31DB9/2020\|EPI_ISL_646197\|20 | ···················· | ···················· | ···················· | ···················· | ···················· | ···················· |
| R | 475 hCoV-19/England/ALDP-B23F96/2020\|EPI_ISL_646198\|20 | ···················· | ···················· | ···················· | ···················· | ···················· | ···················· |
| R | 476 hCoV-19/England/ALDP-B23CAE/2020\|EPI_ISL_646199\|20 | ···················· | ·A·················· | ···················· | ···················· | ···················· | ··A················· |
| R | 477 hCoV-19/England/CAMC-B22FA6/2020\|EPI_ISL_647407\|20 | ···················· | ···················· | ···················· | ···················· | ···················· | ···················· |
| R | 478 hCoV-19/England/CAMC-B22B39/2020\|EPI_ISL_647408\|20 | ···················· | ···················· | ···················· | ···················· | ···················· | ···················· |
| R | 479 hCoV-19/England/CAMC-B229F3/2020\|EPI_ISL_647409\|20 | ···················· | ···················· | ···················· | ···················· | ···················· | ···················· |
| R | 480 hCoV-19/England/CAMC-B22E21/2020\|EPI_ISL_647410\|20 | ···················· | ···················· | ···················· | ···················· | ···················· | ···················· |
| R | 481 hCoV-19/England/CAMC-B216A2/2020\|EPI_ISL_647411\|20 | ···················· | ···················· | ···················· | ···················· | ···················· | ···················· |
| R | 482 hCoV-19/England/CAMC-B22A87/2020\|EPI_ISL_647413\|20 | ···················· | ···················· | ···················· | ···················· | ···················· | ···················· |
| R | 483 hCoV-19/England/CAMC-B219E5/2020\|EPI_ISL_647414\|20 | ···················· | ···················· | ···················· | ···················· | ···················· | ···················· |
| R | 484 hCoV-19/England/CAMC-B22EE5/2020\|EPI_ISL_647415\|20 | ···················· | ···················· | ···················· | ···················· | ···················· | ···················· |
| R | 485 hCoV-19/England/CAMC-B214F3/2020\|EPI_ISL_647416\|20 | ···················· | ···················· | ···················· | ···················· | ···················· | ···················· |
| R | 486 hCoV-19/England/CAMC-B2138D/2020\|EPI_ISL_647417\|20 | ···················· | ·A·················· | ···················· | ···················· | ···················· | ··A················· |
| R | 487 hCoV-19/England/CAMC-B22D06/2020\|EPI_ISL_647418\|20 | ···················· | ···················· | ···················· | ···················· | ···················· | ···················· |
| R | 488 hCoV-19/England/CAMC-B218AC/2020\|EPI_ISL_647419\|20 | ···················· | ···················· | ···················· | ···················· | ···················· | ···················· |
| R | 489 hCoV-19/England/CAMC-B22D7F/2020\|EPI_ISL_647420\|20 | ···················· | ···················· | ···················· | ···················· | ···················· | ···················· |
| R | 490 hCoV-19/England/CAMC-B22832/2020\|EPI_ISL_647425\|20 | ···················· | ···················· | ···················· | ···················· | ···················· | ···················· |
| R | 491 hCoV-19/England/CAMC-B2160C/2020\|EPI_ISL_647426\|20 | ···················· | ···················· | ···················· | ···················· | ···················· | ···················· |
| R | 492 hCoV-19/England/CAMC-B218E8/2020\|EPI_ISL_647427\|20 | ···················· | ·A·················· | ···················· | ···················· | ···················· | ··A················· |
| R | 493 hCoV-19/Singapore/1384/2020\|EPI_ISL_648738\|2020-07 | ···················· | ···················· | ···················· | ···················· | ···················· | ···················· |
| R | 494 hCoV-19/Singapore/1385/2020\|EPI_ISL_648739\|2020-07 | ···················· | ···················· | ···················· | ···················· | ···················· | ···················· |
| R | 495 hCoV-19/Singapore/1386/2020\|EPI_ISL_648740\|2020-07 | ···················· | ···················· | ···················· | ···················· | ···················· | ···················· |
| R | 496 hCoV-19/Singapore/1387/2020\|EPI_ISL_648741\|2020-07 | ···················· | ···················· | ···················· | ···················· | ···················· | ···················· |
| R | 497 hCoV-19/Singapore/1259/2020\|EPI_ISL_648742\|2020-04 | ···················· | ···················· | ···················· | ···················· | ···················· | ···················· |
| R | 498 hCoV-19/Singapore/1409/2020\|EPI_ISL_648743\|2020-05 | ···················· | ···················· | ···················· | ···················· | ···················· | ···················· |
| R | 499 hCoV-19/Singapore/1260/2020\|EPI_ISL_648744\|2020-05 | ···················· | ···················· | ···················· | ···················· | ···················· | ···················· |
| R | 500 hCoV-19/Singapore/1261/2020\|EPI_ISL_648745\|2020-05 | ···················· | ···················· | ···················· | ···················· | ···················· | ···················· |
| R | 501 hCoV-19/Singapore/1262/2020\|EPI_ISL_648746\|2020-05 | ···················· | ···················· | ···················· | ···················· | ···················· | ···················· |
| R | 502 hCoV-19/Singapore/1263/2020\|EPI_ISL_648747\|2020-05 | ···················· | ···················· | ···················· | ···················· | ···················· | ···················· |
| R | 503 hCoV-19/Singapore/1264/2020\|EPI_ISL_648748\|2020-05 | ···················· | ···················· | ···················· | ···················· | ···················· | ···················· |
| R | 504 hCoV-19/Singapore/1265/2020\|EPI_ISL_648749\|2020-05 | ···················· | ···················· | ···················· | ···················· | ···················· | ···················· |
| R | 505 hCoV-19/Singapore/1266/2020\|EPI_ISL_648750\|2020-05 | ···················· | ···················· | ···················· | ···················· | ···················· | ···················· |
| R | 506 hCoV-19/Singapore/1267/2020\|EPI_ISL_648751\|2020-05 | ···················· | ···················· | ···················· | ···················· | ···················· | ···················· |
| R | 507 hCoV-19/Singapore/1270/2020\|EPI_ISL_648754\|2020-05 | ···················· | ···················· | ···················· | ···················· | ···················· | ···················· |
| R | 508 hCoV-19/Singapore/1271/2020\|EPI_ISL_648755\|2020-05 | ···················· | ···················· | ···················· | ···················· | ···················· | ···················· |
| R | 509 hCoV-19/Singapore/1272/2020\|EPI_ISL_648756\|2020-05 | ···················· | ···················· | ···················· | ···················· | ···················· | ···················· |
| R | 510 hCoV-19/Singapore/1273/2020\|EPI_ISL_648757\|2020-05 | ···················· | ···················· | ···················· | ···················· | ···················· | ···················· |
| R | 511 hCoV-19/Singapore/1274/2020\|EPI_ISL_648758\|2020-05 | ···················· | ···················· | ···················· | ···················· | ···················· | ···················· |
| R | 512 hCoV-19/Singapore/885nan/2020\|EPI_ISL_648759\|2020- | ···················· | ···················· | ···················· | ···················· | ···················· | ···················· |
| R | 514 hCoV-19/Australia/VIC12445/2020\|EPI_ISL_641291\|202 | ···················· | ···················· | ···················· | ···················· | ···················· | ···················· |
| R | 515 hCoV-19/Australia/VIC12840/2020\|EPI_ISL_641293\|202 | ···················· | ···················· | ···················· | ···················· | ···················· | ···················· |
| R | 516 hCoV-19/Australia/VIC11684/2020\|EPI_ISL_641294\|202 | ···················· | ···················· | ···················· | ···················· | ···················· | ···················· |
| R | 517 hCoV-19/Australia/VIC16682/2020\|EPI_ISL_641295\|202 | ···················· | ···················· | ···················· | ···················· | ···················· | ···················· |
| R | 518 hCoV-19/Australia/VIC12739/2020\|EPI_ISL_641296\|202 | ···················· | ···················· | ···················· | ···················· | ···················· | ···················· |
| R | 519 hCoV-19/Australia/VIC12892/2020\|EPI_ISL_641297\|202 | ···················· | ···················· | ···················· | ···················· | ···················· | ···················· |
| R | 520 hCoV-19/Australia/VIC12659/2020\|EPI_ISL_641298\|202 | ···················· | ···················· | ···················· | ···················· | ···················· | ···················· |
| R | 521 hCoV-19/Australia/VIC13277/2020\|EPI_ISL_641299\|202 | ···················· | ···················· | ···················· | ···················· | ···················· | ···················· |
| R | 522 hCoV-19/England/QEUH-B10916/2020\|EPI_ISL_642512\|20 | ···················· | ···················· | ···················· | ···················· | ···················· | ···················· |
| R | 523 hCoV-19/England/QEUH-B1012D/2020\|EPI_ISL_642513\|20 | ···················· | ···················· | ···················· | ···················· | ···················· | ···················· |
| R | 524 hCoV-19/England/QEUH-B0EDB5/2020\|EPI_ISL_642514\|20 | ···················· | ···················· | ···················· | ···················· | ···················· | ···················· |
| R | 525 hCoV-19/England/QEUH-B12C39/2020\|EPI_ISL_642515\|20 | ···················· | ···················· | ···················· | ···················· | ···················· | ···················· |
| R | 526 hCoV-19/England/QEUH-B12ECA/2020\|EPI_ISL_642516\|20 | ···················· | ···················· | ···················· | ···················· | ···················· | ···················· |
| R | 527 hCoV-19/England/QEUH-B14350/2020\|EPI_ISL_642517\|20 | ···················· | ···················· | ···················· | ···················· | ···················· | ···················· |
| R | 528 hCoV-19/England/QEUH-B11D91/2020\|EPI_ISL_642518\|20 | ···················· | ···················· | ···················· | ···················· | ···················· | ···················· |
| R | 529 hCoV-19/England/QEUH-B100F3/2020\|EPI_ISL_642519\|20 | ···················· | ···················· | ···················· | ···················· | ···················· | ···················· |
| R | 530 hCoV-19/England/QEUH-B12DEB/2020\|EPI_ISL_642520\|20 | ···················· | ···················· | ···················· | ···················· | ···················· | ···················· |
| R | 531 hCoV-19/England/QEUH-B12422/2020\|EPI_ISL_642521\|20 | ···················· | ···················· | ···················· | ···················· | ···················· | ···················· |
| R | 532 hCoV-19/England/QEUH-B12A6C/2020\|EPI_ISL_642522\|20 | ···················· | ·A·················· | ···················· | ···················· | ···················· | ···················· |
| R | 533 hCoV-19/England/QEUH-B120D3/2020\|EPI_ISL_642527\|20 | ···················· | ···················· | ···················· | ···················· | ···················· | ···················· |
| R | 534 hCoV-19/England/CAMC-B09260/2020\|EPI_ISL_643843\|20 | ···················· | ···················· | ···················· | ···················· | ···················· | ···················· |
| R | 535 hCoV-19/England/CAMC-B083F5/2020\|EPI_ISL_643844\|20 | ···················· | ···················· | ···················· | ···················· | ···················· | ···················· |
| R | 536 hCoV-19/England/CAMC-B07226/2020\|EPI_ISL_643845\|20 | ···················· | ···················· | ···················· | ···················· | ···················· | ···················· |
| R | 537 hCoV-19/England/CAMC-B07FB6/2020\|EPI_ISL_643846\|20 | ···················· | ···················· | ···················· | ···················· | ···················· | ···················· |
| R | 538 hCoV-19/England/CAMC-B088C9/2020\|EPI_ISL_643847\|20 | ···················· | ···················· | ···················· | ···················· | ···················· | ···················· |
| R | 539 hCoV-19/England/CAMC-B0838C/2020\|EPI_ISL_643848\|20 | ···················· | ···················· | ···················· | ···················· | ···················· | ···················· |
| R | 540 hCoV-19/England/CAMC-B07C64/2020\|EPI_ISL_643849\|20 | ···················· | ···················· | ···················· | ···················· | ···················· | ···················· |
| R | 541 hCoV-19/England/CAMC-B08805/2020\|EPI_ISL_643850\|20 | ···················· | ···················· | ···················· | ···················· | ···················· | ···················· |
| R | 542 hCoV-19/England/CAMC-B0729F/2020\|EPI_ISL_643851\|20 | ···················· | ···················· | ···················· | ···················· | ···················· | ···················· |
| R | 543 hCoV-19/England/CAMC-B0746C/2020\|EPI_ISL_643852\|20 | ···················· | ···················· | ···················· | ···················· | ···················· | ···················· |
| R | 544 hCoV-19/England/CAMC-B08173/2020\|EPI_ISL_643858\|20 | ···················· | ···················· | ···················· | ···················· | ···················· | ···················· |
| R | 545 hCoV-19/England/CAMC-B07402/2020\|EPI_ISL_643859\|20 | ···················· | ···················· | ···················· | ···················· | ···················· | ···················· |
| R | 546 hCoV-19/England/ALDP-B1B8C3/2020\|EPI_ISL_646000\|20 | ···················· | ···················· | ···················· | ···················· | ···················· | ···················· |
| R | 547 hCoV-19/England/ALDP-B1A76D/2020\|EPI_ISL_646001\|20 | ···················· | ···················· | ···················· | ···················· | ···················· | ···················· |
| R | 548 hCoV-19/England/ALDP-B1AC22/2020\|EPI_ISL_646002\|20 | ···················· | ···················· | ···················· | ···················· | ···················· | ···················· |
| R | 549 hCoV-19/England/ALDP-B1F768/2020\|EPI_ISL_646003\|20 | ···················· | ···················· | ···················· | ···················· | ···················· | ···················· |
| R | 550 hCoV-19/England/ALDP-B1ACC8/2020\|EPI_ISL_646004\|20 | ···················· | ···················· | ···················· | ···················· | ···················· | ···················· |
| R | 551 hCoV-19/England/ALDP-B1B82D/2020\|EPI_ISL_646005\|20 | ···················· | ···················· | ···················· | ···················· | ···················· | ···················· |
| R | 552 hCoV-19/England/ALDP-B1F5E6/2020\|EPI_ISL_646006\|20 | ···················· | ···················· | ···················· | ···················· | ···················· | ···················· |

| | | | | | | | |
|---|---|---|---|---|---|---|---|
| R | 553 hCoV-19/England/ALDP-B1BD2E/2020\|EPI_ISL_646007\|20 | ···················· | ···················· | ···················· | ···················· | ···················· | ···················· |
| R | 554 hCoV-19/England/ALDP-B1F795/2020\|EPI_ISL_646008\|20 | ···················· | ···················· | ···················· | ···················· | ···················· | ···················· |
| R | 555 hCoV-19/England/ALDP-B1AC40/2020\|EPI_ISL_646010\|20 | ···················· | ···················· | ···················· | ···················· | ···················· | ···················· |
| R | 556 hCoV-19/England/ALDP-B1ACAA/2020\|EPI_ISL_646011\|20 | ···················· | ···················· | ···················· | ···················· | ···················· | ···················· |
| R | 557 hCoV-19/England/ALDP-B1FAAB/2020\|EPI_ISL_646012\|20 | ···················· | ···················· | ···················· | ···················· | ···················· | ···················· |
| R | 558 hCoV-19/England/ALDP-B1AD3E/2020\|EPI_ISL_646013\|20 | ···················· | ···················· | ···················· | ···················· | ···················· | ···················· |
| R | 559 hCoV-19/England/CAMC-B21E6E/2020\|EPI_ISL_646014\|20 | ···················· | ·A·················· | ···················· | ···················· | ···················· | ···A················ |
| R | 560 hCoV-19/England/ALDP-B1DABC/2020\|EPI_ISL_646015\|20 | ···················· | ·A·················· | ···················· | ···················· | ···················· | ···················· |
| R | 561 hCoV-19/England/ALDP-B1AED1/2020\|EPI_ISL_646016\|20 | ···················· | ···················· | ···················· | ···················· | ···················· | ···················· |
| R | 562 hCoV-19/England/CAMC-B221CE/2020\|EPI_ISL_646017\|20 | ···················· | ···················· | ···················· | ···················· | ···················· | ···················· |
| R | 563 hCoV-19/England/CAMC-B223E6/2020\|EPI_ISL_646018\|20 | ···················· | ···················· | ···················· | ···················· | ···················· | ···················· |
| R | 564 hCoV-19/England/ALDP-B1F874/2020\|EPI_ISL_646019\|20 | ···················· | ···················· | ···················· | ···················· | ···················· | ···················· |
| R | 565 hCoV-19/England/ALDP-B1A369/2020\|EPI_ISL_646021\|20 | ···················· | ···················· | ···················· | ···················· | ···················· | ···················· |
| R | 566 hCoV-19/England/CAMC-B21FE3/2020\|EPI_ISL_646022\|20 | ···················· | ···················· | ···················· | ···················· | ···················· | ···················· |
| R | 567 hCoV-19/England/ALDP-B1C15E/2020\|EPI_ISL_646023\|20 | ···················· | ···················· | ···················· | ···················· | ···················· | ···················· |
| R | 568 hCoV-19/England/ALDP-B1A30F/2020\|EPI_ISL_646024\|20 | ···················· | ···················· | ···················· | ···················· | ···················· | ···················· |
| R | 569 hCoV-19/England/ALDP-B1AD7A/2020\|EPI_ISL_646026\|20 | ···················· | ···················· | ···················· | ···················· | ···················· | ···················· |
| R | 570 hCoV-19/England/CAMC-B22568/2020\|EPI_ISL_646027\|20 | ···················· | ···················· | ···················· | ···················· | ···················· | ···················· |
| R | 571 hCoV-19/England/CAMC-B2201C/2020\|EPI_ISL_646028\|20 | ···················· | ···················· | ···················· | ···················· | ···················· | ···················· |
| R | 572 hCoV-19/England/ALDP-B1FA7E/2020\|EPI_ISL_646029\|20 | ···················· | ···················· | ···················· | ···················· | ···················· | ···················· |
| R | 573 hCoV-19/England/CAMC-B2229E/2020\|EPI_ISL_646030\|20 | ···················· | ···················· | ···················· | ···················· | ···················· | ···················· |
| R | 574 hCoV-19/England/CAMC-B21FC5/2020\|EPI_ISL_646032\|20 | ···················· | ···················· | ···················· | ···················· | ···················· | ···················· |
| R | 575 hCoV-19/England/ALDP-B1AE1D/2020\|EPI_ISL_646033\|20 | ···················· | ···················· | ···················· | ···················· | ···················· | ···················· |
| R | 576 hCoV-19/England/CAMC-B22119/2020\|EPI_ISL_646034\|20 | ···················· | ···················· | ···················· | ···················· | ···················· | ···················· |
| R | 577 hCoV-19/England/CAMC-B22085/2020\|EPI_ISL_646035\|20 | ···················· | ···················· | ···················· | ···················· | ···················· | ···················· |
| R | 578 hCoV-19/England/ALDP-B1AD98/2020\|EPI_ISL_646036\|20 | ···················· | ···················· | ···················· | ···················· | ···················· | ···················· |
| R | 579 hCoV-19/England/ALDP-B1F7D1/2020\|EPI_ISL_646037\|20 | ···················· | ···················· | ···················· | ···················· | ···················· | ···················· |
| R | 580 hCoV-19/England/ALDP-B1F63E/2020\|EPI_ISL_646038\|20 | ···················· | ···················· | ···················· | ···················· | ···················· | ···················· |
| R | 581 hCoV-19/England/CAMC-B225FF/2020\|EPI_ISL_646039\|20 | ···················· | ···················· | ···················· | ···················· | ···················· | ···················· |
| R | 582 hCoV-19/England/ALDP-B1C455/2020\|EPI_ISL_646040\|20 | ···················· | ···················· | ···················· | ···················· | ···················· | ···················· |
| R | 583 hCoV-19/England/CAMC-B2203A/2020\|EPI_ISL_646041\|20 | ···················· | ···················· | ···················· | ···················· | ···················· | ···················· |
| R | 584 hCoV-19/England/ALDP-B1D551/2020\|EPI_ISL_646042\|20 | ···················· | ···················· | ···················· | ···················· | ···················· | ···················· |
| R | 585 hCoV-19/England/ALDP-B1C060/2020\|EPI_ISL_646043\|20 | ···················· | ···················· | ···················· | ···················· | ···················· | ···················· |
| R | 586 hCoV-19/England/ALDP-B1AE4A/2020\|EPI_ISL_646044\|20 | ···················· | ···················· | ···················· | ···················· | ···················· | ···················· |
| R | 587 hCoV-19/England/CAMC-B21D61/2020\|EPI_ISL_646046\|20 | ···················· | ···················· | ···················· | ···················· | ······T············· | ···················· |
| R | 588 hCoV-19/England/CAMC-B225D1/2020\|EPI_ISL_646047\|20 | ···················· | ·A·················· | ···················· | ···················· | ···················· | ···A················ |
| R | 589 hCoV-19/England/ALDP-B1AE68/2020\|EPI_ISL_646048\|20 | ···················· | ···················· | ···················· | ···················· | ···················· | ···················· |
| R | 590 hCoV-19/England/ALDP-B1F8A1/2020\|EPI_ISL_646049\|20 | ···················· | ···················· | ···················· | ···················· | ···················· | ···················· |
| R | 591 hCoV-19/England/ALDP-B1A3C3/2020\|EPI_ISL_646050\|20 | ···················· | ···················· | ···················· | ···················· | ···················· | ···················· |
| R | 592 hCoV-19/England/CAMC-B2235F/2020\|EPI_ISL_646052\|20 | ···················· | ···················· | ···················· | ···················· | ···················· | ···················· |
| R | 593 hCoV-19/England/ALDP-B1C0CA/2020\|EPI_ISL_646053\|20 | ···················· | ···················· | ···················· | ···················· | ···················· | ···A················ |
| R | 594 hCoV-19/England/ALDP-B1B1AA/2020\|EPI_ISL_646054\|20 | ···················· | ···················· | ···················· | ···················· | ···················· | ···················· |
| R | 595 hCoV-19/England/ALDP-B1DB7D/2020\|EPI_ISL_646055\|20 | ···················· | ···················· | ···················· | ···················· | ···················· | ···················· |
| R | 596 hCoV-19/England/CAMC-B220EF/2020\|EPI_ISL_646056\|20 | ···················· | ···················· | ···················· | ···················· | ···················· | ···················· |
| R | 597 hCoV-19/England/CAMC-B223F5/2020\|EPI_ISL_646058\|20 | ···················· | ···················· | ···················· | ···················· | ······T············· | ···················· |
| R | 598 hCoV-19/England/ALDP-B1A2E4/2020\|EPI_ISL_646059\|20 | ···················· | ···················· | ···················· | ···················· | ···················· | ···················· |
| R | 599 hCoV-19/England/CAMC-B228D8/2020\|EPI_ISL_647308\|20 | ···················· | ···················· | ···················· | ···················· | ···················· | ···················· |
| R | 600 hCoV-19/England/CAMC-B22D24/2020\|EPI_ISL_647310\|20 | ···················· | ···················· | ···················· | ···················· | ···················· | ···················· |
| R | 601 hCoV-19/England/CAMC-B226CF/2020\|EPI_ISL_647311\|20 | ···················· | ···················· | ···················· | ···················· | ······T············· | ···················· |
| R | 602 hCoV-19/England/CAMC-B22C36/2020\|EPI_ISL_647312\|20 | ···················· | ···················· | ···················· | ···················· | ···················· | ···················· |
| R | 603 hCoV-19/England/CAMC-B2295D/2020\|EPI_ISL_647313\|20 | ···················· | ···················· | ···················· | ···················· | ···················· | ···················· |
| R | 604 hCoV-19/England/CAMC-B2288D/2020\|EPI_ISL_647314\|20 | ···················· | ···················· | ···················· | ···················· | ···················· | ···················· |
| R | 605 hCoV-19/England/CAMC-B213E7/2020\|EPI_ISL_647315\|20 | ···················· | ···················· | ···················· | ···················· | ···················· | ···················· |
| R | 606 hCoV-19/England/CAMC-B2162A/2020\|EPI_ISL_647316\|20 | ···················· | ···················· | ···················· | ···················· | ···················· | ···················· |
| R | 607 hCoV-19/England/CAMC-B22D60/2020\|EPI_ISL_647319\|20 | ···················· | ···················· | ···················· | ···················· | ···················· | ···················· |
| R | 608 hCoV-19/England/CAMC-B2196D/2020\|EPI_ISL_647320\|20 | ···················· | ···················· | ···················· | ···················· | ···················· | ···················· |
| R | 609 hCoV-19/England/CAMC-B21E13/2020\|EPI_ISL_647321\|20 | ···················· | ·A·················· | ···················· | ···················· | ···················· | ···A················ |
| R | 610 hCoV-19/England/CAMC-B21A97/2020\|EPI_ISL_647323\|20 | ···················· | ···················· | ···················· | ···················· | ···················· | ···················· |
| R | 611 hCoV-19/England/CAMC-B21DF8/2020\|EPI_ISL_647324\|20 | ···················· | ···················· | ···················· | ···················· | ···················· | ···················· |
| R | 612 hCoV-19/England/CAMC-B219A9/2020\|EPI_ISL_647325\|20 | ···················· | ···················· | ···················· | ···················· | ···················· | ···················· |
| R | 613 hCoV-19/England/CAMC-B21F4D/2020\|EPI_ISL_647327\|20 | ···················· | ···················· | ···················· | ···················· | ···················· | ···················· |
| R | 614 hCoV-19/England/CAMC-B21903/2020\|EPI_ISL_647328\|20 | ···················· | ···················· | ···················· | ···················· | ···················· | ···················· |
| R | 615 hCoV-19/England/CAMC-B22E6D/2020\|EPI_ISL_647330\|20 | ···················· | ···················· | ···················· | ···················· | ···················· | ···················· |
| R | 616 hCoV-19/England/CAMC-B2199A/2020\|EPI_ISL_647331\|20 | ···················· | ···················· | ···················· | ···················· | ···················· | ···················· |
| R | 617 hCoV-19/England/CAMC-B21CFB/2020\|EPI_ISL_647332\|20 | ···················· | ···················· | ···················· | ···················· | ···················· | ···················· |
| R | 618 hCoV-19/England/CAMC-B213AB/2020\|EPI_ISL_647333\|20 | ···················· | ···················· | ···················· | ···················· | ···················· | ···················· |
| R | 619 hCoV-19/England/CAMC-B21AD3/2020\|EPI_ISL_647334\|20 | ···················· | ···················· | ···················· | ···················· | ···················· | ···A················ |
| R | 620 hCoV-19/England/CAMC-B22C90/2020\|EPI_ISL_647335\|20 | ···················· | ···················· | ···················· | ···················· | ···················· | ···················· |
| R | 621 hCoV-19/England/CAMC-B21727/2020\|EPI_ISL_647336\|20 | ···················· | ···················· | ···················· | ···················· | ···················· | ···················· |
| R | 622 hCoV-19/England/CAMC-B227DB/2020\|EPI_ISL_647337\|20 | ···················· | ···················· | ···················· | ···················· | ···················· | ···················· |
| R | 623 hCoV-19/England/CAMC-B228F6/2020\|EPI_ISL_647339\|20 | ···················· | ···················· | ···················· | ···················· | ···················· | ···················· |
| R | 624 hCoV-19/England/CAMC-B22B84/2020\|EPI_ISL_647340\|20 | ···················· | ···················· | ···················· | ···················· | ···················· | ···················· |
| R | 625 hCoV-19/England/CAMC-B2287E/2020\|EPI_ISL_647342\|20 | ···················· | ···················· | ···················· | ···················· | ···················· | ···················· |
| R | 626 hCoV-19/England/CAMC-B21D8F/2020\|EPI_ISL_647343\|20 | ···················· | ···················· | ···················· | ···················· | ···················· | ···················· |
| R | 627 hCoV-19/England/CAMC-B221BF/2020\|EPI_ISL_647344\|20 | ···················· | ···················· | ···················· | ···················· | ···················· | ···················· |
| R | 628 hCoV-19/England/CAMC-B23303/2020\|EPI_ISL_647345\|20 | ···················· | ···················· | ···················· | ···················· | ···················· | ···················· |
| R | 629 hCoV-19/England/CAMC-B22EF4/2020\|EPI_ISL_647346\|20 | ···················· | ···················· | ···················· | ···················· | ···················· | ···················· |
| R | 630 hCoV-19/England/CAMC-B21C82/2020\|EPI_ISL_647347\|20 | ···················· | ···················· | ···················· | ···················· | ····T··············· | ···················· |
| R | 631 hCoV-19/England/CAMC-B229C6/2020\|EPI_ISL_647348\|20 | ···················· | ·A·················· | ···················· | ···················· | ···················· | ···················· |
| R | 632 hCoV-19/England/CAMC-B2102C/2020\|EPI_ISL_647350\|20 | ···················· | ···················· | ···················· | ···················· | ···················· | ···················· |

<table>
<tr><td>R</td><td>633 hCoV-19/England/CAMC-B21666/2020|EPI_ISL_647351|20</td><td>····················</td><td>····················</td><td>·····················</td><td>····················</td><td>····················</td><td>····················</td></tr>
<tr><td>R</td><td>634 hCoV-19/England/CAMC-B2101D/2020|EPI_ISL_647352|20</td><td>····················</td><td>····················</td><td>·····················</td><td>····················</td><td>····················</td><td>····················</td></tr>
<tr><td>R</td><td>635 hCoV-19/England/CAMC-B21815/2020|EPI_ISL_647353|20</td><td>····················</td><td>····················</td><td>·····················</td><td>····················</td><td>····················</td><td>····················</td></tr>
<tr><td>R</td><td>636 hCoV-19/England/CAMC-B229E4/2020|EPI_ISL_647357|20</td><td>····················</td><td>····················</td><td>·····················</td><td>····················</td><td>····················</td><td>····················</td></tr>
<tr><td>R</td><td>637 hCoV-19/England/CAMC-B22726/2020|EPI_ISL_647358|20</td><td>····················</td><td>····················</td><td>·····················</td><td>····················</td><td>····················</td><td>····················</td></tr>
<tr><td>R</td><td>638 hCoV-19/England/CAMC-B22C72/2020|EPI_ISL_647360|20</td><td>····················</td><td>····················</td><td>·····················</td><td>····················</td><td>····················</td><td>····················</td></tr>
<tr><td>R</td><td>639 hCoV-19/England/CAMC-B21930/2020|EPI_ISL_647361|20</td><td>····················</td><td>····················</td><td>·····················</td><td>····················</td><td>····················</td><td>····················</td></tr>
<tr><td>R</td><td>640 hCoV-19/England/CAMC-B21CCE/2020|EPI_ISL_647362|20</td><td>····················</td><td>····················</td><td>·····················</td><td>····················</td><td>····T···············</td><td>····················</td></tr>
<tr><td>R</td><td>641 hCoV-19/England/CAMC-B21E5F/2020|EPI_ISL_647363|20</td><td>····················</td><td>····················</td><td>·····················</td><td>····················</td><td>····················</td><td>····················</td></tr>
<tr><td>R</td><td>642 hCoV-19/England/CAMC-B22A3C/2020|EPI_ISL_647364|20</td><td>····················</td><td>····················</td><td>·····················</td><td>····················</td><td>····················</td><td>····················</td></tr>
<tr><td>R</td><td>643 hCoV-19/England/CAMC-B21D70/2020|EPI_ISL_647365|20</td><td>····················</td><td>····················</td><td>·····················</td><td>····················</td><td>····················</td><td>····················</td></tr>
<tr><td>R</td><td>644 hCoV-19/England/CAMC-B22B1B/2020|EPI_ISL_647367|20</td><td>····················</td><td>····················</td><td>·····················</td><td>····················</td><td>····················</td><td>····················</td></tr>
<tr><td>R</td><td>645 hCoV-19/England/CAMC-B21C46/2020|EPI_ISL_647369|20</td><td>····················</td><td>····················</td><td>·····················</td><td>····················</td><td>····················</td><td>····················</td></tr>
<tr><td>R</td><td>646 hCoV-19/England/CAMC-B22FE2/2020|EPI_ISL_647371|20</td><td>····················</td><td>····················</td><td>·····················</td><td>····················</td><td>·····T··············</td><td>····················</td></tr>
<tr><td>R</td><td>647 hCoV-19/England/CAMC-B22841/2020|EPI_ISL_647372|20</td><td>····················</td><td>····················</td><td>·····················</td><td>····················</td><td>····················</td><td>····················</td></tr>
<tr><td>R</td><td>648 hCoV-19/England/CAMC-B2298A/2020|EPI_ISL_647374|20</td><td>····················</td><td>····················</td><td>·····················</td><td>····················</td><td>····················</td><td>····················</td></tr>
<tr><td>R</td><td>649 hCoV-19/England/CAMC-B22C63/2020|EPI_ISL_647375|20</td><td>····················</td><td>····················</td><td>·····················</td><td>····················</td><td>····················</td><td>····················</td></tr>
<tr><td>R</td><td>650 hCoV-19/England/CAMC-B21C19/2020|EPI_ISL_647376|20</td><td>····················</td><td>·A··················</td><td>·····················</td><td>····················</td><td>····················</td><td>··A·················</td></tr>
<tr><td>R</td><td>651 hCoV-19/England/CAMC-B22FC4/2020|EPI_ISL_647377|20</td><td>····················</td><td>····················</td><td>·····················</td><td>····················</td><td>····················</td><td>····················</td></tr>
<tr><td>R</td><td>652 hCoV-19/England/CAMC-B2103B/2020|EPI_ISL_647379|20</td><td>····················</td><td>····················</td><td>·····················</td><td>····················</td><td>····················</td><td>····················</td></tr>
<tr><td>R</td><td>653 hCoV-19/England/CAMC-B22E4F/2020|EPI_ISL_647380|20</td><td>····················</td><td>····················</td><td>·····················</td><td>····················</td><td>····················</td><td>····················</td></tr>
<tr><td>R</td><td>654 hCoV-19/England/CAMC-B215B4/2020|EPI_ISL_647381|20</td><td>····················</td><td>····················</td><td>·····················</td><td>····················</td><td>····················</td><td>····················</td></tr>
<tr><td>R</td><td>655 hCoV-19/England/CAMC-B21068/2020|EPI_ISL_647382|20</td><td>····················</td><td>·A··················</td><td>·····················</td><td>····················</td><td>····················</td><td>··A·················</td></tr>
<tr><td>R</td><td>656 hCoV-19/England/CAMC-B2104A/2020|EPI_ISL_647383|20</td><td>····················</td><td>·A··················</td><td>·····················</td><td>····················</td><td>····················</td><td>··A·················</td></tr>
<tr><td>R</td><td>657 hCoV-19/England/CAMC-B22665/2020|EPI_ISL_647385|20</td><td>····················</td><td>····················</td><td>·····················</td><td>····················</td><td>····················</td><td>····················</td></tr>
<tr><td>R</td><td>658 hCoV-19/England/CAMC-B217CD/2020|EPI_ISL_647386|20</td><td>····················</td><td>····················</td><td>·····················</td><td>····················</td><td>····················</td><td>····················</td></tr>
<tr><td>R</td><td>659 hCoV-19/England/CAMC-B22B66/2020|EPI_ISL_647387|20</td><td>····················</td><td>····················</td><td>·····················</td><td>····················</td><td>····················</td><td>····················</td></tr>
<tr><td>R</td><td>660 hCoV-19/England/CAMC-B232F7/2020|EPI_ISL_647388|20</td><td>····················</td><td>····················</td><td>·····················</td><td>····················</td><td>····················</td><td>····················</td></tr>
<tr><td>R</td><td>661 hCoV-19/England/CAMC-B2245C/2020|EPI_ISL_647400|20</td><td>····················</td><td>····················</td><td>·····················</td><td>····················</td><td>····················</td><td>····················</td></tr>
<tr><td>R</td><td>662 hCoV-19/England/CAMC-B22E12/2020|EPI_ISL_647401|20</td><td>····················</td><td>····················</td><td>·····················</td><td>····················</td><td>····················</td><td>····················</td></tr>
<tr><td>R</td><td>663 hCoV-19/England/CAMC-B2139C/2020|EPI_ISL_647402|20</td><td>····················</td><td>····················</td><td>·····················</td><td>····················</td><td>····················</td><td>····················</td></tr>
<tr><td>R</td><td>664 hCoV-19/England/CAMC-B22647/2020|EPI_ISL_647403|20</td><td>····················</td><td>····················</td><td>·····················</td><td>····················</td><td>····················</td><td>····················</td></tr>
<tr><td>R</td><td>665 hCoV-19/England/CAMC-B22F97/2020|EPI_ISL_647404|20</td><td>····················</td><td>····················</td><td>·····················</td><td>····················</td><td>·····T··············</td><td>····················</td></tr>
<tr><td>R</td><td>666 hCoV-19/England/CAMC-B2286F/2020|EPI_ISL_647405|20</td><td>····················</td><td>····················</td><td>·····················</td><td>····················</td><td>····················</td><td>····················</td></tr>
<tr><td>R</td><td>667 hCoV-19/England/CAMC-B21A2E/2020|EPI_ISL_647406|20</td><td>····················</td><td>····················</td><td>·····················</td><td>····················</td><td>····················</td><td>····················</td></tr>
<tr><td>R</td><td>668 hCoV-19/England/CAMC-B22B75/2020|EPI_ISL_647421|20</td><td>····················</td><td>····················</td><td>·····················</td><td>····················</td><td>····················</td><td>····················</td></tr>
<tr><td>R</td><td>669 hCoV-19/Singapore/1223/2020|EPI_ISL_648639|2020-04</td><td>····················</td><td>····················</td><td>·····················</td><td>····················</td><td>····················</td><td>····················</td></tr>
<tr><td>R</td><td>670 hCoV-19/Singapore/1224/2020|EPI_ISL_648640|2020-04</td><td>····················</td><td>····················</td><td>·····················</td><td>····················</td><td>····················</td><td>····················</td></tr>
<tr><td>R</td><td>671 hCoV-19/Singapore/1225/2020|EPI_ISL_648641|2020-04</td><td>····················</td><td>····················</td><td>·····················</td><td>····················</td><td>····················</td><td>····················</td></tr>
<tr><td>R</td><td>672 hCoV-19/Singapore/1226/2020|EPI_ISL_648642|2020-04</td><td>····················</td><td>····················</td><td>·····················</td><td>····················</td><td>····················</td><td>····················</td></tr>
<tr><td>R</td><td>673 hCoV-19/Singapore/1227/2020|EPI_ISL_648643|2020-04</td><td>····················</td><td>····················</td><td>·····················</td><td>····················</td><td>····················</td><td>····················</td></tr>
<tr><td>R</td><td>674 hCoV-19/Singapore/1228/2020|EPI_ISL_648644|2020-04</td><td>····················</td><td>····················</td><td>·····················</td><td>····················</td><td>····················</td><td>····················</td></tr>
<tr><td>R</td><td>675 hCoV-19/Singapore/1229/2020|EPI_ISL_648645|2020-04</td><td>····················</td><td>····················</td><td>·····················</td><td>····················</td><td>····················</td><td>····················</td></tr>
<tr><td>R</td><td>676 hCoV-19/Singapore/1230/2020|EPI_ISL_648646|2020-04</td><td>····················</td><td>····················</td><td>·····················</td><td>····················</td><td>····················</td><td>····················</td></tr>
<tr><td>R</td><td>677 hCoV-19/Singapore/1231/2020|EPI_ISL_648647|2020-04</td><td>····················</td><td>····················</td><td>·····················</td><td>····················</td><td>····················</td><td>····················</td></tr>
<tr><td>R</td><td>678 hCoV-19/Singapore/415nan/2020|EPI_ISL_648648|2020-</td><td>····················</td><td>····················</td><td>·····················</td><td>····················</td><td>····················</td><td>····················</td></tr>
<tr><td>R</td><td>679 hCoV-19/Singapore/1232/2020|EPI_ISL_648649|2020-04</td><td>····················</td><td>····················</td><td>·····················</td><td>····················</td><td>····················</td><td>····················</td></tr>
<tr><td>R</td><td>680 hCoV-19/Singapore/1233/2020|EPI_ISL_648650|2020-04</td><td>····················</td><td>····················</td><td>·····················</td><td>····················</td><td>····················</td><td>····················</td></tr>
<tr><td>R</td><td>681 hCoV-19/Singapore/1234/2020|EPI_ISL_648651|2020-04</td><td>····················</td><td>····················</td><td>·····················</td><td>····················</td><td>····················</td><td>····················</td></tr>
<tr><td>R</td><td>682 hCoV-19/Singapore/1235/2020|EPI_ISL_648652|2020-04</td><td>····················</td><td>····················</td><td>·····················</td><td>····················</td><td>····················</td><td>····················</td></tr>
<tr><td>R</td><td>683 hCoV-19/Singapore/1236/2020|EPI_ISL_648653|2020-04</td><td>····················</td><td>····················</td><td>·····················</td><td>····················</td><td>····················</td><td>····················</td></tr>
<tr><td>R</td><td>684 hCoV-19/Singapore/1237/2020|EPI_ISL_648654|2020-04</td><td>····················</td><td>····················</td><td>·····················</td><td>····················</td><td>····················</td><td>····················</td></tr>
<tr><td>R</td><td>685 hCoV-19/Singapore/1238/2020|EPI_ISL_648655|2020-04</td><td>····················</td><td>····················</td><td>·····················</td><td>····················</td><td>····················</td><td>····················</td></tr>
<tr><td>R</td><td>686 hCoV-19/Singapore/1239/2020|EPI_ISL_648656|2020-04</td><td>····················</td><td>····················</td><td>·····················</td><td>····················</td><td>····················</td><td>····················</td></tr>
<tr><td>R</td><td>687 hCoV-19/Singapore/1240/2020|EPI_ISL_648657|2020-04</td><td>····················</td><td>····················</td><td>·····················</td><td>····················</td><td>····················</td><td>····················</td></tr>
<tr><td>R</td><td>688 hCoV-19/Singapore/1241/2020|EPI_ISL_648658|2020-04</td><td>····················</td><td>····················</td><td>·····················</td><td>····················</td><td>····················</td><td>····················</td></tr>
<tr><td>R</td><td>689 hCoV-19/Singapore/1242/2020|EPI_ISL_648659|2020-04</td><td>····················</td><td>····················</td><td>·····················</td><td>····················</td><td>····················</td><td>····················</td></tr>
<tr><td>R</td><td>690 hCoV-19/Singapore/1243/2020|EPI_ISL_648660|2020-04</td><td>····················</td><td>····················</td><td>·····················</td><td>····················</td><td>····················</td><td>····················</td></tr>
<tr><td>R</td><td>691 hCoV-19/Singapore/1408/2020|EPI_ISL_648661|2020-05</td><td>····················</td><td>····················</td><td>·····················</td><td>····················</td><td>····················</td><td>····················</td></tr>
<tr><td>R</td><td>692 hCoV-19/Singapore/1244/2020|EPI_ISL_648662|2020-05</td><td>····················</td><td>····················</td><td>·····················</td><td>····················</td><td>····················</td><td>····················</td></tr>
<tr><td>R</td><td>693 hCoV-19/Singapore/1245/2020|EPI_ISL_648663|2020-05</td><td>····················</td><td>····················</td><td>·····················</td><td>····················</td><td>····················</td><td>····················</td></tr>
<tr><td>R</td><td>694 hCoV-19/Singapore/1246/2020|EPI_ISL_648664|2020-05</td><td>····················</td><td>····················</td><td>·····················</td><td>····················</td><td>····················</td><td>····················</td></tr>
<tr><td>R</td><td>695 hCoV-19/Singapore/1247/2020|EPI_ISL_648665|2020-05</td><td>····················</td><td>····················</td><td>·····················</td><td>····················</td><td>····················</td><td>····················</td></tr>
<tr><td>R</td><td>696 hCoV-19/Singapore/1248/2020|EPI_ISL_648666|2020-05</td><td>····················</td><td>····················</td><td>·····················</td><td>····················</td><td>····················</td><td>····················</td></tr>
<tr><td>R</td><td>697 hCoV-19/Singapore/1249/2020|EPI_ISL_648667|2020-05</td><td>····················</td><td>····················</td><td>·····················</td><td>····················</td><td>····················</td><td>····················</td></tr>
<tr><td>R</td><td>698 hCoV-19/Singapore/1250/2020|EPI_ISL_648668|2020-05</td><td>····················</td><td>····················</td><td>·····················</td><td>····················</td><td>····················</td><td>····················</td></tr>
<tr><td>R</td><td>699 hCoV-19/Singapore/1251/2020|EPI_ISL_648669|2020-05</td><td>····················</td><td>····················</td><td>·····················</td><td>····················</td><td>····················</td><td>····················</td></tr>
<tr><td>R</td><td>700 hCoV-19/Singapore/1252/2020|EPI_ISL_648670|2020-05</td><td>····················</td><td>····················</td><td>·····················</td><td>····················</td><td>····················</td><td>····················</td></tr>
<tr><td>R</td><td>701 hCoV-19/Singapore/1253/2020|EPI_ISL_648671|2020-05</td><td>····················</td><td>····················</td><td>·····················</td><td>····················</td><td>····················</td><td>····················</td></tr>
<tr><td>R</td><td>702 hCoV-19/Singapore/1254/2020|EPI_ISL_648672|2020-05</td><td>····················</td><td>····················</td><td>·····················</td><td>····················</td><td>····················</td><td>····················</td></tr>
<tr><td>R</td><td>703 hCoV-19/Singapore/1255/2020|EPI_ISL_648673|2020-05</td><td>····················</td><td>····················</td><td>·····················</td><td>····················</td><td>····················</td><td>····················</td></tr>
<tr><td>R</td><td>704 hCoV-19/Singapore/1256/2020|EPI_ISL_648674|2020-05</td><td>····················</td><td>····················</td><td>·····················</td><td>····················</td><td>····················</td><td>····················</td></tr>
<tr><td>R</td><td>705 hCoV-19/Singapore/1257/2020|EPI_ISL_648675|2020-05</td><td>····················</td><td>····················</td><td>·····················</td><td>····················</td><td>····················</td><td>····················</td></tr>
<tr><td>R</td><td>706 hCoV-19/Singapore/1258/2020|EPI_ISL_648676|2020-05</td><td>····················</td><td>····················</td><td>·····················</td><td>····················</td><td>····················</td><td>····················</td></tr>
<tr><td>R</td><td>707 hCoV-19/Argentina/argenTAG-1252C02E/2020|EPI_ISL_6</td><td>····················</td><td>····················</td><td>·····················</td><td>····················</td><td>····················</td><td>····················</td></tr>
<tr><td>R</td><td>708 hCoV-19/Singapore/1332/2020|EPI_ISL_648678|2020-07</td><td>····················</td><td>····················</td><td>·····················</td><td>····················</td><td>····················</td><td>····················</td></tr>
<tr><td>R</td><td>709 hCoV-19/Singapore/702nan/2020|EPI_ISL_648679|2020-</td><td>····················</td><td>····················</td><td>·····················</td><td>····················</td><td>····················</td><td>····················</td></tr>
<tr><td>R</td><td>710 hCoV-19/Singapore/695nan/2020|EPI_ISL_648680|2020-</td><td>····················</td><td>····················</td><td>·····················</td><td>····················</td><td>····················</td><td>····················</td></tr>
<tr><td>R</td><td>711 hCoV-19/Singapore/1411/2020|EPI_ISL_648681|2020-07</td><td>····················</td><td>····················</td><td>·····················</td><td>····················</td><td>····················</td><td>····················</td></tr>
<tr><td>R</td><td>712 hCoV-19/Singapore/1333/2020|EPI_ISL_648682|2020-06</td><td>····················</td><td>····················</td><td>·····················</td><td>····················</td><td>····················</td><td>····················</td></tr>
</table>

| | | | | | | | |
|---|---|---|---|---|---|---|---|
| R | 713 hCoV-19/Singapore/1334/2020\|EPI_ISL_648683\|2020-06 | .................... | .................... | .................... | .................... | .................... | .................... |
| R | 714 hCoV-19/Singapore/1335/2020\|EPI_ISL_648684\|2020-06 | .................... | .................... | .................... | .................... | .................... | .................... |
| R | 715 hCoV-19/Singapore/1336/2020\|EPI_ISL_648685\|2020-06 | .................... | .................... | .................... | .................... | .................... | .................... |
| R | 716 hCoV-19/Singapore/1338/2020\|EPI_ISL_648687\|2020-06 | .................... | .................... | .................... | .................... | .................... | .................... |
| R | 717 hCoV-19/Singapore/1339/2020\|EPI_ISL_648688\|2020-06 | .................... | .................... | .................... | .................... | .................... | .................... |
| R | 718 hCoV-19/Singapore/1340/2020\|EPI_ISL_648689\|2020-06 | .................... | .................... | .................... | .................... | .................... | .................... |
| R | 719 hCoV-19/Singapore/1341/2020\|EPI_ISL_648690\|2020-06 | .................... | .................... | .................... | .................... | .................... | .................... |
| R | 720 hCoV-19/Singapore/1342/2020\|EPI_ISL_648691\|2020-06 | .................... | .................... | .................... | .................... | .................... | .................... |
| R | 721 hCoV-19/Singapore/1343/2020\|EPI_ISL_648692\|2020-06 | .................... | .................... | .................... | .................... | .................... | .................... |
| R | 722 hCoV-19/Singapore/1344/2020\|EPI_ISL_648693\|2020-06 | .................... | .................... | .................... | .................... | .................... | .................... |
| R | 723 hCoV-19/Singapore/1345/2020\|EPI_ISL_648694\|2020-06 | .................... | .................... | .................... | .................... | .................... | .................... |
| R | 724 hCoV-19/Singapore/1346/2020\|EPI_ISL_648695\|2020-06 | .................... | .................... | .................... | .................... | .................... | .................... |
| R | 725 hCoV-19/Singapore/1347/2020\|EPI_ISL_648696\|2020-06 | .................... | .................... | .................... | .................... | .................... | .................... |
| R | 726 hCoV-19/Singapore/1348/2020\|EPI_ISL_648697\|2020-06 | .................... | .................... | .................... | .................... | .................... | .................... |
| R | 727 hCoV-19/Singapore/707nan/2020\|EPI_ISL_648698\|2020- | .................... | .................... | .................... | .................... | .................... | .................... |
| R | 728 hCoV-19/Singapore/1349/2020\|EPI_ISL_648699\|2020-06 | .................... | .................... | .................... | .................... | .................... | .................... |
| R | 729 hCoV-19/Singapore/1350/2020\|EPI_ISL_648700\|2020-06 | .................... | .................... | .................... | .................... | .................... | .................... |
| R | 730 hCoV-19/Singapore/1351/2020\|EPI_ISL_648701\|2020-06 | .................... | .................... | .................... | .................... | .................... | .................... |
| R | 731 hCoV-19/Singapore/599nan/2020\|EPI_ISL_648702\|2020- | .................... | .................... | .................... | .................... | .................... | .................... |
| R | 732 hCoV-19/Singapore/1352/2020\|EPI_ISL_648703\|2020-06 | .................... | .................... | .................... | .................... | .................... | .................... |
| R | 733 hCoV-19/Singapore/1353/2020\|EPI_ISL_648704\|2020-06 | .................... | .................... | .................... | .................... | .................... | .................... |
| R | 734 hCoV-19/Singapore/1354/2020\|EPI_ISL_648705\|2020-06 | .................... | .................... | .................... | .................... | .................... | .................... |
| R | 735 hCoV-19/Singapore/1355/2020\|EPI_ISL_648706\|2020-06 | .................... | .................... | .................... | .................... | .................... | .................... |
| R | 736 hCoV-19/Singapore/1356/2020\|EPI_ISL_648707\|2020-06 | .................... | .................... | .................... | .................... | .................... | .................... |
| R | 737 hCoV-19/Singapore/1357/2020\|EPI_ISL_648708\|2020-06 | .................... | .................... | .................... | .................... | .................... | .................... |
| R | 738 hCoV-19/Singapore/1358/2020\|EPI_ISL_648709\|2020-06 | .................... | .................... | .................... | .................... | .................... | .................... |
| R | 739 hCoV-19/Singapore/1359/2020\|EPI_ISL_648710\|2020-06 | .................... | .................... | .................... | .................... | .................... | .................... |
| R | 740 hCoV-19/Singapore/1360/2020\|EPI_ISL_648711\|2020-06 | .................... | .................... | .................... | .................... | .................... | .................... |
| R | 741 hCoV-19/Singapore/1361/2020\|EPI_ISL_648712\|2020-06 | .................... | .................... | .................... | .................... | .................... | .................... |
| R | 742 hCoV-19/Singapore/1362/2020\|EPI_ISL_648713\|2020-06 | .................... | .................... | .................... | .................... | .................... | .................... |
| R | 743 hCoV-19/Singapore/1363/2020\|EPI_ISL_648714\|2020-06 | .................... | .................... | .................... | .................... | .................... | .................... |
| R | 744 hCoV-19/Singapore/1364/2020\|EPI_ISL_648715\|2020-06 | .................... | .................... | .................... | .................... | .................... | .................... |
| R | 745 hCoV-19/Singapore/591nan/2020\|EPI_ISL_648716\|2020- | .................... | .................... | .................... | .................... | .................... | .................... |
| R | 746 hCoV-19/Singapore/1365/2020\|EPI_ISL_648717\|2020-06 | .................... | .................... | .................... | .................... | .................... | .................... |
| R | 747 hCoV-19/Singapore/1366/2020\|EPI_ISL_648718\|2020-06 | .................... | .................... | .................... | .................... | .................... | .................... |
| R | 748 hCoV-19/Singapore/1367/2020\|EPI_ISL_648719\|2020-06 | .................... | .................... | .................... | .................... | .................... | .................... |
| R | 749 hCoV-19/Singapore/1368/2020\|EPI_ISL_648720\|2020-06 | .................... | .................... | .................... | .................... | .................... | .................... |
| R | 750 hCoV-19/Singapore/1369/2020\|EPI_ISL_648721\|2020-06 | .................... | .................... | .................... | .................... | .................... | .................... |
| R | 751 hCoV-19/Singapore/1371/2020\|EPI_ISL_648722\|2020-06 | .................... | .................... | .................... | .................... | .................... | .................... |
| R | 752 hCoV-19/Singapore/1372/2020\|EPI_ISL_648723\|2020-07 | .................... | .................... | .................... | .................... | .................... | .................... |
| R | 753 hCoV-19/Singapore/667nan/2020\|EPI_ISL_648724\|2020- | .................... | .................... | .................... | .................... | .................... | .................... |
| R | 754 hCoV-19/Singapore/1373/2020\|EPI_ISL_648725\|2020-07 | .................... | .................... | .................... | .................... | .................... | .................... |
| R | 755 hCoV-19/Singapore/600nan/2020\|EPI_ISL_648726\|2020- | .................... | .................... | .................... | .................... | .................... | .................... |
| R | 756 hCoV-19/Singapore/1374/2020\|EPI_ISL_648727\|2020-07 | .................... | .................... | .................... | .................... | .................... | .................... |
| R | 757 hCoV-19/Singapore/1375/2020\|EPI_ISL_648728\|2020-07 | .................... | .................... | .................... | .................... | .................... | .................... |
| R | 758 hCoV-19/Singapore/1376/2020\|EPI_ISL_648729\|2020-07 | .................... | .................... | .................... | .................... | .................... | .................... |
| R | 759 hCoV-19/Singapore/1377/2020\|EPI_ISL_648730\|2020-07 | .................... | .................... | .................... | .................... | .................... | .................... |
| R | 760 hCoV-19/Singapore/822nan/2020\|EPI_ISL_648731\|2020- | .................... | .................... | .................... | .................... | .................... | .................... |
| R | 761 hCoV-19/Singapore/1378/2020\|EPI_ISL_648732\|2020-07 | .................... | .................... | .................... | .................... | .................... | .................... |
| R | 762 hCoV-19/Singapore/1379/2020\|EPI_ISL_648733\|2020-07 | .................... | .................... | .................... | .................... | .................... | .................... |
| R | 763 hCoV-19/Singapore/1380/2020\|EPI_ISL_648734\|2020-07 | .................... | .................... | .................... | .................... | .................... | .................... |
| R | 764 hCoV-19/Singapore/1381/2020\|EPI_ISL_648735\|2020-07 | .................... | .................... | .................... | .................... | .................... | .................... |
| R | 765 hCoV-19/Singapore/1382/2020\|EPI_ISL_648736\|2020-07 | .................... | .................... | .................... | .................... | .................... | .................... |
| R | 766 hCoV-19/Singapore/1383/2020\|EPI_ISL_648737\|2020-07 | .................... | .................... | .................... | .................... | .................... | .................... |
| R | 767 hCoV-19/Singapore/1268/2020\|EPI_ISL_648752\|2020-05 | .................... | .................... | .................... | .................... | .................... | .................... |
| R | 768 hCoV-19/Singapore/1269/2020\|EPI_ISL_648753\|2020-05 | .................... | .................... | .................... | .................... | .................... | .................... |
| R | 769 hCoV-19/Australia/VIC15984/2020\|EPI_ISL_641104\|202 | .................... | .................... | .................... | .................... | .................... | .................... |
| R | 770 hCoV-19/Australia/VIC13166/2020\|EPI_ISL_641105\|202 | .................... | .................... | .................... | .................... | .................... | .................... |
| R | 771 hCoV-19/Australia/VIC11729/2020\|EPI_ISL_641107\|202 | .................... | .................... | .................... | .................... | .................... | .................... |
| R | 772 hCoV-19/Australia/VIC12136/2020\|EPI_ISL_641108\|202 | .................... | .................... | .................... | .................... | .................... | .................... |
| R | 773 hCoV-19/Australia/VIC16229/2020\|EPI_ISL_641109\|202 | .................... | .................... | .................... | .................... | .................... | .................... |
| R | 774 hCoV-19/Australia/VIC12567/2020\|EPI_ISL_641111\|202 | .................... | .................... | .................... | .................... | .................... | .................... |
| R | 776 hCoV-19/Australia/VIC15954/2020\|EPI_ISL_641113\|202 | .................... | .................... | .................... | .................... | .................... | .................... |
| R | 777 hCoV-19/Australia/VIC13060/2020\|EPI_ISL_641114\|202 | .................... | .................... | .................... | .................... | .................... | .................... |
| R | 778 hCoV-19/Australia/VIC16104/2020\|EPI_ISL_641115\|202 | .................... | .................... | .................... | .................... | .................... | .................... |
| R | 779 hCoV-19/Australia/VIC12370/2020\|EPI_ISL_641117\|202 | .................... | .................... | .................... | .................... | .................... | .................... |
| R | 780 hCoV-19/Australia/VIC15962/2020\|EPI_ISL_641118\|202 | .................... | .................... | .................... | .................... | .................... | .................... |
| R | 781 hCoV-19/Australia/VIC12908/2020\|EPI_ISL_641120\|202 | .................... | .................... | .................... | .................... | .................... | .................... |
| R | 782 hCoV-19/Australia/VIC11948/2020\|EPI_ISL_641121\|202 | .................... | .................... | .................... | .................... | .................... | .................... |
| R | 783 hCoV-19/Australia/VIC16522/2020\|EPI_ISL_641122\|202 | .................... | .................... | .................... | .................... | .................... | .................... |
| R | 784 hCoV-19/Australia/VIC16499/2020\|EPI_ISL_641123\|202 | .................... | .................... | .................... | .................... | .................... | .................... |
| R | 785 hCoV-19/Australia/VIC16634/2020\|EPI_ISL_641124\|202 | .................... | .................... | .................... | .................... | .................... | .................... |
| R | 786 hCoV-19/Australia/VIC12187/2020\|EPI_ISL_641125\|202 | .................... | .................... | .................... | .................... | .................... | .................... |
| R | 787 hCoV-19/Australia/VIC12470/2020\|EPI_ISL_641128\|202 | .................... | .................... | .................... | .................... | .................... | .................... |
| R | 788 hCoV-19/Australia/VIC12814/2020\|EPI_ISL_641129\|202 | .................... | .................... | .................... | .................... | .................... | .................... |
| R | 789 hCoV-19/Australia/VIC12403/2020\|EPI_ISL_641130\|202 | .................... | .................... | .................... | .................... | .................... | .................... |
| R | 790 hCoV-19/Australia/VIC12490/2020\|EPI_ISL_641131\|202 | .................... | .................... | .................... | .................... | .................... | .................... |
| R | 791 hCoV-19/Australia/VIC12561/2020\|EPI_ISL_641132\|202 | .................... | .................... | .................... | .................... | .................... | .................... |
| R | 792 hCoV-19/Australia/VIC12135/2020\|EPI_ISL_641133\|202 | .................... | .................... | .................... | .................... | .................... | .................... |
| R | 793 hCoV-19/Australia/VIC16088/2020\|EPI_ISL_641134\|202 | .................... | .................... | .................... | .................... | .................... | .................... |

| | | | | | | | | |
|---|---|---|---|---|---|---|---|---|
| R | 794 hCoV-19/Australia/VIC16211/2020\|EPI_ISL_641135\|202 | .................... | .................... | .................... | .................... | .................... | .................... | |
| R | 795 hCoV-19/Australia/VIC15910/2020\|EPI_ISL_641136\|202 | .................... | .................... | .................... | .................... | .................... | .................... | |
| R | 796 hCoV-19/Australia/VIC16339/2020\|EPI_ISL_641137\|202 | .................... | .................... | .................... | .................... | .................... | .................... | |
| R | 797 hCoV-19/Australia/VIC16417/2020\|EPI_ISL_641138\|202 | .................... | .................... | .................... | .................... | .................... | .................... | |
| R | 798 hCoV-19/Australia/VIC12853/2020\|EPI_ISL_641140\|202 | .................... | .................... | .................... | .................... | .................... | .................... | |
| R | 799 hCoV-19/Australia/VIC15958/2020\|EPI_ISL_641141\|202 | .................... | .................... | .................... | .................... | .................... | .................... | |
| R | 800 hCoV-19/Australia/VIC16138/2020\|EPI_ISL_641142\|202 | .................... | .................... | .................... | .................... | .................... | .................... | |
| R | 801 hCoV-19/Australia/VIC12531/2020\|EPI_ISL_641146\|202 | .................... | .................... | .................... | .................... | .................... | .................... | |
| R | 802 hCoV-19/Australia/VIC12150/2020\|EPI_ISL_641147\|202 | .................... | .................... | .................... | .................... | .................... | .................... | |
| R | 804 hCoV-19/Australia/VIC16102/2020\|EPI_ISL_641149\|202 | .................... | .................... | .................... | .................... | .................... | .................... | |
| R | 805 hCoV-19/Australia/VIC16353/2020\|EPI_ISL_641150\|202 | .................... | .................... | .................... | .................... | .................... | .................... | |
| R | 806 hCoV-19/Australia/VIC13219/2020\|EPI_ISL_641151\|202 | .................... | .................... | .................... | .................... | .................... | .................... | |
| R | 807 hCoV-19/Australia/VIC12697/2020\|EPI_ISL_641152\|202 | .................... | .................... | .................... | .................... | .................... | .................... | |
| R | 808 hCoV-19/Australia/VIC16119/2020\|EPI_ISL_641153\|202 | .................... | .................... | .................... | .................... | .................... | .................... | |
| R | 809 hCoV-19/Australia/VIC12080/2020\|EPI_ISL_641154\|202 | .................... | .................... | .................... | .................... | .................... | .................... | |
| R | 810 hCoV-19/Australia/VIC16067/2020\|EPI_ISL_641155\|202 | .................... | .................... | .................... | .................... | .................... | .................... | |
| R | 811 hCoV-19/Australia/VIC12568/2020\|EPI_ISL_641156\|202 | .................... | .................... | .................... | .................... | .................... | .................... | |
| R | 812 hCoV-19/Australia/VIC16427/2020\|EPI_ISL_641157\|202 | .................... | .................... | .................... | .................... | .................... | .................... | |
| R | 813 hCoV-19/Australia/VIC16645/2020\|EPI_ISL_641158\|202 | .................... | .................... | .................... | .................... | .................... | .................... | |
| R | 814 hCoV-19/Australia/VIC12514/2020\|EPI_ISL_641159\|202 | .................... | .................... | .................... | .................... | .................... | .................... | |
| R | 815 hCoV-19/Australia/VIC12139/2020\|EPI_ISL_641160\|202 | .................... | .................... | .................... | .................... | NNN................. | .................... | 1 |
| R | 816 hCoV-19/Australia/VIC11671/2020\|EPI_ISL_641161\|202 | .................... | .................... | .................... | .................... | .................... | .................... | |
| R | 817 hCoV-19/Australia/VIC16432/2020\|EPI_ISL_641162\|202 | .................... | .................... | .................... | .................... | .................... | .................... | |
| R | 818 hCoV-19/Australia/VIC16155/2020\|EPI_ISL_641164\|202 | .................... | .................... | .................... | .................... | .................... | .................... | |
| R | 819 hCoV-19/Australia/VIC16492/2020\|EPI_ISL_641167\|202 | .................... | .................... | .................... | .................... | .................... | .................... | |
| R | 820 hCoV-19/Australia/VIC12434/2020\|EPI_ISL_641168\|202 | .................... | .................... | .................... | .................... | .................... | .................... | |
| R | 821 hCoV-19/Australia/VIC16288/2020\|EPI_ISL_641200\|202 | .................... | .................... | .................... | .................... | .................... | .................... | |
| R | 822 hCoV-19/Australia/VIC12515/2020\|EPI_ISL_641201\|202 | .................... | .................... | .................... | .................... | .................... | .................... | |
| R | 823 hCoV-19/England/QEUH-B115E4/2020\|EPI_ISL_642413\|20 | .................... | .................... | .................... | .................... | .................... | .................... | |
| R | 824 hCoV-19/England/QEUH-B11D46/2020\|EPI_ISL_642414\|20 | .................... | .................... | .................... | .................... | .................... | .................... | |
| R | 825 hCoV-19/England/QEUH-B10E71/2020\|EPI_ISL_642415\|20 | .................... | .................... | .................... | .................... | .................... | .................... | |
| R | 826 hCoV-19/England/QEUH-B130B4/2020\|EPI_ISL_642416\|20 | .................... | .................... | .................... | .................... | .................... | .................... | |
| R | 827 hCoV-19/England/QEUH-B11739/2020\|EPI_ISL_642417\|20 | .................... | .................... | .................... | .................... | .................... | .................... | |
| R | 828 hCoV-19/England/QEUH-B1054F/2020\|EPI_ISL_642418\|20 | .................... | .................... | .................... | .................... | .................... | .................... | |
| R | 829 hCoV-19/England/QEUH-B121EF/2020\|EPI_ISL_642420\|20 | .................... | .................... | .................... | .................... | .................... | .................... | |
| R | 830 hCoV-19/England/QEUH-B11414/2020\|EPI_ISL_642422\|20 | .................... | .................... | .................... | .................... | .................... | .................... | |
| R | 831 hCoV-19/England/QEUH-B131DF/2020\|EPI_ISL_642435\|20 | .................... | .................... | .................... | .................... | .................... | .................... | |
| R | 832 hCoV-19/England/QEUH-B131B1/2020\|EPI_ISL_642437\|20 | .................... | .................... | .................... | .................... | .................... | .................... | |
| R | 833 hCoV-19/England/QEUH-B13193/2020\|EPI_ISL_642439\|20 | .................... | .................... | .................... | .................... | .................... | .................... | |
| R | 834 hCoV-19/England/QEUH-B11A12/2020\|EPI_ISL_642440\|20 | .................... | .................... | .................... | .................... | .................... | .................... | |
| R | 835 hCoV-19/England/QEUH-B11B97/2020\|EPI_ISL_642441\|20 | .................... | .................... | .................... | .................... | .................... | .................... | |
| R | 836 hCoV-19/England/QEUH-B11A03/2020\|EPI_ISL_642442\|20 | .................... | .................... | .................... | .................... | .................... | .................... | |
| R | 837 hCoV-19/England/QEUH-B11B4C/2020\|EPI_ISL_642444\|20 | .................... | .................... | .................... | .................... | .................... | .................... | |
| R | 838 hCoV-19/England/QEUH-B1157B/2020\|EPI_ISL_642445\|20 | .................... | .................... | .................... | .................... | .................... | .................... | |
| R | 839 hCoV-19/England/QEUH-B11942/2020\|EPI_ISL_642446\|20 | .................... | .................... | .................... | .................... | .................... | .................... | |
| R | 840 hCoV-19/England/QEUH-B11CFE/2020\|EPI_ISL_642447\|20 | .................... | .................... | .................... | .................... | .................... | .................... | |
| R | 841 hCoV-19/England/QEUH-B12932/2020\|EPI_ISL_642448\|20 | .................... | .................... | .................... | .................... | .................... | .................... | |
| R | 842 hCoV-19/England/QEUH-B11C49/2020\|EPI_ISL_642449\|20 | .................... | .A.................. | .................... | .................... | .................... | .................... | |
| R | 843 hCoV-19/England/QEUH-B11915/2020\|EPI_ISL_642451\|20 | .................... | .................... | .................... | .................... | .................... | .................... | |
| R | 844 hCoV-19/England/QEUH-B1147E/2020\|EPI_ISL_642452\|20 | .................... | .................... | .................... | .................... | .................... | .................... | |
| R | 845 hCoV-19/England/QEUH-B11863/2020\|EPI_ISL_642454\|20 | .................... | .................... | .................... | .................... | .................... | .................... | |
| R | 846 hCoV-19/England/QEUH-B135F1/2020\|EPI_ISL_642455\|20 | .................... | .................... | .................... | .................... | .................... | .................... | |
| R | 847 hCoV-19/England/QEUH-B1270B/2020\|EPI_ISL_642456\|20 | .................... | .................... | .................... | .................... | .................... | .................... | |
| R | 848 hCoV-19/England/QEUH-B11B00/2020\|EPI_ISL_642457\|20 | .................... | .................... | .................... | .................... | .................... | .................... | |
| R | 849 hCoV-19/England/QEUH-B114E7/2020\|EPI_ISL_642459\|20 | .................... | .A.................. | .................... | .................... | .................... | .................... | |
| R | 850 hCoV-19/England/QEUH-B11696/2020\|EPI_ISL_642460\|20 | .................... | .................... | .................... | .................... | .................... | .................... | |
| R | 851 hCoV-19/England/QEUH-B11B5B/2020\|EPI_ISL_642461\|20 | .................... | .................... | .................... | .................... | .................... | .................... | |
| R | 852 hCoV-19/England/QEUH-B1056D/2020\|EPI_ISL_642462\|20 | .................... | .................... | .................... | .................... | .................... | .................... | |
| R | 853 hCoV-19/England/QEUH-B13166/2020\|EPI_ISL_642463\|20 | .................... | .................... | .................... | .................... | .................... | .................... | |
| R | 854 hCoV-19/England/QEUH-B1164B/2020\|EPI_ISL_642464\|20 | .................... | .................... | .................... | .................... | .................... | .................... | |
| R | 855 hCoV-19/England/QEUH-B11C58/2020\|EPI_ISL_642465\|20 | .................... | .................... | .................... | .................... | .................... | .................... | |
| R | 856 hCoV-19/England/QEUH-B10512/2020\|EPI_ISL_642466\|20 | .................... | .................... | .................... | .................... | .................... | .................... | |
| R | 857 hCoV-19/England/QEUH-B1047F/2020\|EPI_ISL_642467\|20 | .................... | .................... | .................... | .................... | .................... | .................... | |
| R | 858 hCoV-19/England/QEUH-B11E52/2020\|EPI_ISL_642468\|20 | .................... | .................... | .................... | .................... | .................... | .................... | |
| R | 859 hCoV-19/England/QEUH-B12246/2020\|EPI_ISL_642469\|20 | .................... | .................... | .................... | .................... | .................... | .................... | |
| R | 860 hCoV-19/England/QEUH-B12783/2020\|EPI_ISL_642470\|20 | .................... | .................... | .................... | .................... | .................... | .................... | |
| R | 861 hCoV-19/England/QEUH-B12158/2020\|EPI_ISL_642472\|20 | .................... | .................... | .................... | .................... | .................... | .................... | |
| R | 862 hCoV-19/England/QEUH-B11757/2020\|EPI_ISL_642473\|20 | .................... | .................... | .................... | .................... | .................... | .................... | |
| R | 863 hCoV-19/England/QEUH-B119F7/2020\|EPI_ISL_642474\|20 | .................... | .................... | .................... | .................... | .................... | .................... | |
| R | 864 hCoV-19/England/QEUH-B119CA/2020\|EPI_ISL_642475\|20 | .................... | .................... | .................... | .................... | .................... | .................... | |
| R | 865 hCoV-19/England/QEUH-B11766/2020\|EPI_ISL_642476\|20 | .................... | .................... | .................... | .................... | .................... | .................... | |
| R | 866 hCoV-19/England/QEUH-B130E1/2020\|EPI_ISL_642477\|20 | .................... | .................... | .................... | .................... | .................... | .................... | |
| R | 867 hCoV-19/England/QEUH-B12255/2020\|EPI_ISL_642478\|20 | .................... | .................... | .................... | .................... | .................... | .................... | |
| R | 868 hCoV-19/England/QEUH-B1161E/2020\|EPI_ISL_642481\|20 | .................... | .................... | .................... | .................... | .................... | .................... | |
| R | 869 hCoV-19/England/QEUH-B10F23/2020\|EPI_ISL_642482\|20 | .................... | .................... | .................... | .................... | .................... | .................... | |
| R | 870 hCoV-19/England/QEUH-B12808/2020\|EPI_ISL_642484\|20 | .................... | .................... | .................... | .................... | .................... | .................... | |
| R | 871 hCoV-19/England/QEUH-B11A30/2020\|EPI_ISL_642485\|20 | .................... | .................... | .................... | .................... | .................... | .................... | |
| R | 872 hCoV-19/England/QEUH-B104CA/2020\|EPI_ISL_642486\|20 | .................... | .................... | .................... | .................... | .................... | .................... | |
| R | 873 hCoV-19/England/QEUH-B11793/2020\|EPI_ISL_642488\|20 | .................... | .................... | .................... | .................... | .................... | .................... | |
| R | 874 hCoV-19/England/QEUH-B12EBB/2020\|EPI_ISL_642489\|20 | .................... | .................... | .................... | .................... | .................... | .................... | |

| | | | | | | | |
|---|---|---|---|---|---|---|---|
| R | 875 hCoV-19/England/QEUH-B1260E/2020\|EPI_ISL_642490\|20 | ···················· | ···················· | ····················· | ···················· | ···················· | ···················· |
| R | 876 hCoV-19/England/QEUH-B10C0E/2020\|EPI_ISL_642491\|20 | ···················· | ···················· | ····················· | ···················· | ···················· | ···················· |
| R | 877 hCoV-19/England/QEUH-B12343/2020\|EPI_ISL_642492\|20 | ···················· | ···················· | ····················· | ···················· | ···················· | ···················· |
| R | 878 hCoV-19/England/QEUH-B1460C/2020\|EPI_ISL_642495\|20 | ···················· | ···················· | ····················· | ···················· | ···················· | ···················· |
| R | 879 hCoV-19/England/QEUH-B144D5/2020\|EPI_ISL_642496\|20 | ···················· | ···················· | ····················· | ···················· | ···················· | ···················· |
| R | 880 hCoV-19/England/QEUH-B12B1E/2020\|EPI_ISL_642497\|20 | ···················· | ···················· | ····················· | ···················· | ···················· | ···················· |
| R | 881 hCoV-19/England/QEUH-B1451E/2020\|EPI_ISL_642498\|20 | ···················· | ···················· | ····················· | ···················· | ······T············· | ···················· |
| R | 882 hCoV-19/England/QEUH-B0F1C7/2020\|EPI_ISL_642500\|20 | ···················· | ···················· | ····················· | ···················· | ···················· | ···················· |
| R | 883 hCoV-19/England/QEUH-B12A5D/2020\|EPI_ISL_642501\|20 | ···················· | ···················· | ····················· | ···················· | ···················· | ···················· |
| R | 884 hCoV-19/Wales/QEUH-B1245F/2020\|EPI_ISL_642502\|2020 | ···················· | ···················· | ····················· | ···················· | ···················· | ···················· |
| R | 885 hCoV-19/England/QEUH-B12AE4/2020\|EPI_ISL_642503\|20 | ···················· | ···················· | ····················· | ···················· | ···················· | ···················· |
| R | 886 hCoV-19/England/QEUH-B113EA/2020\|EPI_ISL_642504\|20 | ···················· | ·A·················· | ····················· | ···················· | ···················· | ···················· |
| R | 887 hCoV-19/England/QEUH-B122FB/2020\|EPI_ISL_642507\|20 | ···················· | ···················· | ····················· | ···················· | ···················· | ···················· |
| R | 888 hCoV-19/England/QEUH-B1212B/2020\|EPI_ISL_642508\|20 | ···················· | ···················· | ····················· | ···················· | ···················· | ···················· |
| R | 889 hCoV-19/England/QEUH-B1436F/2020\|EPI_ISL_642509\|20 | ···················· | ···················· | ····················· | ···················· | ···················· | ···················· |
| R | 890 hCoV-19/England/QEUH-B12598/2020\|EPI_ISL_642510\|20 | ···················· | ···················· | ····················· | ···················· | ···················· | ···················· |
| R | 891 hCoV-19/England/QEUH-B12EE8/2020\|EPI_ISL_642511\|20 | ···················· | ···················· | ····················· | ···················· | ···················· | ···················· |
| R | 892 hCoV-19/England/QEUH-B0F1B8/2020\|EPI_ISL_642523\|20 | ···················· | ···················· | ····················· | ···················· | ···················· | ···················· |
| R | 893 hCoV-19/England/QEUH-B12316/2020\|EPI_ISL_642524\|20 | ·····G·············· | ···················· | ····················· | ···················· | ···················· | ···················· |
| R | 894 hCoV-19/England/QEUH-B10925/2020\|EPI_ISL_642525\|20 | ···················· | ···················· | ····················· | ···················· | ···················· | ···················· |
| R | 895 hCoV-19/England/QEUH-B1211C/2020\|EPI_ISL_642526\|20 | ···················· | ···················· | ····················· | ···················· | ···················· | ···················· |
| R | 896 hCoV-19/England/QEUH-B12C0C/2020\|EPI_ISL_642528\|20 | ···················· | ···················· | ····················· | ···················· | ···················· | ···················· |
| R | 897 hCoV-19/England/QEUH-B1210D/2020\|EPI_ISL_642529\|20 | ···················· | ···················· | ····················· | ···················· | ···················· | ···················· |
| R | 898 hCoV-19/England/QEUH-B122CE/2020\|EPI_ISL_642530\|20 | ···················· | ···················· | ····················· | ···················· | ···················· | ···················· |
| R | 899 hCoV-19/England/QEUH-B1201F/2020\|EPI_ISL_642531\|20 | ···················· | ···················· | ····················· | ···················· | ···················· | ···················· |
| R | 900 hCoV-19/England/QEUH-B11292/2020\|EPI_ISL_642532\|20 | ···················· | ···················· | ····················· | ···················· | ···················· | ···················· |
| R | 901 hCoV-19/England/QEUH-B11256/2020\|EPI_ISL_642533\|20 | ···················· | ···················· | ····················· | ···················· | ···················· | ···················· |
| R | 902 hCoV-19/England/CAMC-B08674/2020\|EPI_ISL_643744\|20 | ···················· | ···················· | ····················· | ···················· | ···················· | ···················· |
| R | 903 hCoV-19/England/CAMC-B07F10/2020\|EPI_ISL_643745\|20 | ···················· | ···················· | ····················· | ···················· | ···················· | ···················· |
| R | 904 hCoV-19/England/CAMC-B07A3D/2020\|EPI_ISL_643746\|20 | ···················· | ···················· | ····················· | ···················· | ···················· | ···················· |
| R | 905 hCoV-19/England/CAMC-B07851/2020\|EPI_ISL_643747\|20 | ···················· | ·A·················· | ····················· | ···················· | ···················· | ···A················ |
| R | 906 hCoV-19/England/CAMC-B0928E/2020\|EPI_ISL_643748\|20 | ···················· | ···················· | ····················· | ···················· | ···················· | ···················· |
| R | 907 hCoV-19/England/CAMC-B08AF0/2020\|EPI_ISL_643751\|20 | ···················· | ···················· | ····················· | ···················· | ···················· | ···················· |
| R | 908 hCoV-19/England/CAMC-B07B85/2020\|EPI_ISL_643753\|20 | ···················· | ···················· | ····················· | ···················· | ···················· | ···················· |
| R | 909 hCoV-19/England/CAMC-B07BEF/2020\|EPI_ISL_643754\|20 | ···················· | ···················· | ····················· | ···················· | ···················· | ···················· |
| R | 910 hCoV-19/England/CAMC-B092D9/2020\|EPI_ISL_643766\|20 | ···················· | ···················· | ····················· | ···················· | ···················· | ···················· |
| R | 911 hCoV-19/England/CAMC-B089E4/2020\|EPI_ISL_643767\|20 | ···················· | ···················· | ····················· | ···················· | ···················· | ···················· |
| R | 912 hCoV-19/England/CAMC-B0887E/2020\|EPI_ISL_643768\|20 | ···················· | ···················· | ····················· | ···················· | ···················· | ···················· |
| R | 913 hCoV-19/England/CAMC-B074F3/2020\|EPI_ISL_643769\|20 | ···················· | ···················· | ····················· | ···················· | ···················· | ···················· |
| R | 914 hCoV-19/England/CAMC-B08823/2020\|EPI_ISL_643770\|20 | ···················· | ···················· | ····················· | ···················· | ···················· | ···················· |
| R | 915 hCoV-19/England/CAMC-B08A0F/2020\|EPI_ISL_643771\|20 | ···················· | ···················· | ····················· | ···················· | ···················· | ···················· |
| R | 916 hCoV-19/England/CAMC-B08B2A/2020\|EPI_ISL_643772\|20 | ···················· | ···················· | ····················· | ···················· | ···················· | ···················· |
| R | 917 hCoV-19/England/CAMC-B085D1/2020\|EPI_ISL_643773\|20 | ···················· | ···················· | ····················· | ···················· | ···················· | ···················· |
| R | 918 hCoV-19/England/CAMC-B079B8/2020\|EPI_ISL_643774\|20 | ···················· | ···················· | ····················· | ···················· | ···················· | ···················· |
| R | 919 hCoV-19/England/CAMC-B0748A/2020\|EPI_ISL_643775\|20 | ···················· | ···················· | ····················· | ···················· | ···················· | ···················· |
| R | 920 hCoV-19/England/CAMC-B07068/2020\|EPI_ISL_643776\|20 | ···················· | ···················· | ····················· | ···················· | ···················· | ···················· |
| R | 921 hCoV-19/England/CAMC-B08629/2020\|EPI_ISL_643777\|20 | ···················· | ···················· | ····················· | ···················· | ···················· | ···················· |
| R | 922 hCoV-19/England/CAMC-B073E7/2020\|EPI_ISL_643778\|20 | ···················· | ···················· | ····················· | ···················· | ······T············· | ···················· |
| R | 923 hCoV-19/England/CAMC-B0761B/2020\|EPI_ISL_643779\|20 | ···················· | ···················· | ····················· | ···················· | ···················· | ···················· |
| R | 924 hCoV-19/England/CAMC-B07D16/2020\|EPI_ISL_643780\|20 | ···················· | ···················· | ····················· | ···················· | ···················· | ···················· |
| R | 925 hCoV-19/England/CAMC-B0789D/2020\|EPI_ISL_643781\|20 | ···················· | ···················· | ····················· | ···················· | ···················· | ···················· |
| R | 926 hCoV-19/England/CAMC-B0754B/2020\|EPI_ISL_643782\|20 | ···················· | ···················· | ····················· | ···················· | ···················· | ···················· |
| R | 927 hCoV-19/England/CAMC-B08717/2020\|EPI_ISL_643785\|20 | ···················· | ···················· | ····················· | ···················· | ···················· | ···················· |
| R | 928 hCoV-19/England/CAMC-B08665/2020\|EPI_ISL_643786\|20 | ···················· | ···················· | ····················· | ···················· | ···················· | ···················· |
| R | 929 hCoV-19/England/CAMC-B07095/2020\|EPI_ISL_643787\|20 | ···················· | ···················· | ····················· | ···················· | ···················· | ···················· |
| R | 930 hCoV-19/England/CAMC-B07CDD/2020\|EPI_ISL_643788\|20 | ···················· | ···················· | ····················· | ···················· | ···················· | ···················· |
| R | 931 hCoV-19/England/CAMC-B08C27/2020\|EPI_ISL_643789\|20 | ···················· | ···················· | ····················· | ···················· | ···················· | ···················· |
| R | 932 hCoV-19/England/CAMC-B07A2E/2020\|EPI_ISL_643790\|20 | ···················· | ···················· | ····················· | ···················· | ···················· | ···················· |
| R | 933 hCoV-19/England/CAMC-B08832/2020\|EPI_ISL_643791\|20 | ···················· | ···················· | ····················· | ···················· | ···················· | ···················· |
| R | 934 hCoV-19/England/CAMC-B07596/2020\|EPI_ISL_643792\|20 | ···················· | ·A·················· | ····················· | ···················· | ···················· | ···A················ |
| R | 935 hCoV-19/England/CAMC-B08C90/2020\|EPI_ISL_643793\|20 | ···················· | ···················· | ····················· | ···················· | ···················· | ···················· |
| R | 936 hCoV-19/England/CAMC-B07244/2020\|EPI_ISL_643794\|20 | ···················· | ···················· | ····················· | ···················· | ···················· | ···················· |
| R | 937 hCoV-19/England/CAMC-B0750F/2020\|EPI_ISL_643795\|20 | ···················· | ···················· | ····················· | ···················· | ······T············· | ···················· |
| R | 938 hCoV-19/England/CAMC-B08911/2020\|EPI_ISL_643796\|20 | ···················· | ···················· | ····················· | ···················· | ···················· | ···················· |
| R | 939 hCoV-19/England/CAMC-B08A78/2020\|EPI_ISL_643798\|20 | ···················· | ···················· | ····················· | ···················· | ···················· | ···················· |
| R | 940 hCoV-19/England/CAMC-B07271/2020\|EPI_ISL_643799\|20 | ···················· | ···················· | ····················· | ···················· | ···················· | ···················· |
| R | 941 hCoV-19/England/CAMC-B086DE/2020\|EPI_ISL_643800\|20 | ···················· | ···················· | ····················· | ···················· | ···················· | ···················· |
| R | 942 hCoV-19/England/CAMC-B07077/2020\|EPI_ISL_643801\|20 | ···················· | ···················· | ····················· | ···················· | ···················· | ···················· |
| R | 943 hCoV-19/England/CAMC-B070B3/2020\|EPI_ISL_643802\|20 | ···················· | ···················· | ····················· | ···················· | ···················· | ···················· |
| R | 944 hCoV-19/England/CAMC-B08BFD/2020\|EPI_ISL_643803\|20 | ···················· | ···················· | ····················· | ···················· | ···················· | ···················· |
| R | 945 hCoV-19/England/CAMC-B0787F/2020\|EPI_ISL_643804\|20 | ···················· | ···················· | ····················· | ···················· | ···················· | ···················· |
| R | 946 hCoV-19/England/CAMC-B08692/2020\|EPI_ISL_643806\|20 | ···················· | ···················· | ····················· | ···················· | ···················· | ···················· |
| R | 947 hCoV-19/England/CAMC-B08164/2020\|EPI_ISL_643807\|20 | ···················· | ···················· | ····················· | ···················· | ···················· | ···················· |
| R | 948 hCoV-19/England/CAMC-B07860/2020\|EPI_ISL_643808\|20 | ···················· | ···················· | ····················· | ···················· | ···················· | ···················· |
| R | 949 hCoV-19/England/CAMC-B08D15/2020\|EPI_ISL_643809\|20 | ···················· | ···················· | ····················· | ···················· | ···················· | ···················· |
| R | 950 hCoV-19/England/CAMC-B07921/2020\|EPI_ISL_643810\|20 | ···················· | ···················· | ····················· | ···················· | ···················· | ···················· |
| R | 951 hCoV-19/England/CAMC-B07332/2020\|EPI_ISL_643811\|20 | ···················· | ···················· | ····················· | ···················· | ···················· | ···················· |
| R | 952 hCoV-19/England/CAMC-B076C0/2020\|EPI_ISL_643812\|20 | ···················· | ···················· | ····················· | ···················· | ···················· | ···················· |
| R | 953 hCoV-19/England/CAMC-B07824/2020\|EPI_ISL_643813\|20 | ···················· | ···················· | ····················· | ···················· | ···················· | ···················· |
| R | 954 hCoV-19/England/CAMC-B07D25/2020\|EPI_ISL_643816\|20 | ···················· | ···················· | ····················· | ···················· | ···················· | ···················· |

| | | | | | | | | |
|---|---|---|---|---|---|---|---|---|
| R | 955 | hCoV-19/England/CAMC-B07930/2020\|EPI_ISL_643817\|20 | ···················· | ···················· | ····················· | ···················· | ···················· | ···················· |
| R | 956 | hCoV-19/England/CAMC-B088F6/2020\|EPI_ISL_643818\|20 | ···················· | ···················· | ····················· | ···················· | ···················· | ···················· |
| R | 957 | hCoV-19/England/CAMC-B071FC/2020\|EPI_ISL_643819\|20 | ···················· | ···················· | ····················· | ···················· | ···················· | ···················· |
| R | 958 | hCoV-19/England/CAMC-B0752D/2020\|EPI_ISL_643820\|20 | ···················· | ···················· | ····················· | ···················· | ···················· | ···················· |
| R | 959 | hCoV-19/England/CAMC-B0743F/2020\|EPI_ISL_643821\|20 | ···················· | ···················· | ····················· | ···················· | ···················· | ···················· |
| R | 960 | hCoV-19/England/CAMC-B072EA/2020\|EPI_ISL_643822\|20 | ···················· | ···················· | ····················· | ···················· | ···················· | ···················· |
| R | 961 | hCoV-19/England/CAMC-B071A1/2020\|EPI_ISL_643823\|20 | ···················· | ···················· | ····················· | ···················· | ······T············· | ···················· |
| R | 962 | hCoV-19/England/CAMC-B07648/2020\|EPI_ISL_643824\|20 | ···················· | ···················· | ····················· | ···················· | ······T············· | ···················· |
| R | 963 | hCoV-19/England/CAMC-B07736/2020\|EPI_ISL_643825\|20 | ···················· | ···················· | ····················· | ···················· | ···················· | ···················· |
| R | 964 | hCoV-19/England/CAMC-B08C81/2020\|EPI_ISL_643826\|20 | ···················· | ···················· | ····················· | ···················· | ···················· | ···················· |
| R | 965 | hCoV-19/England/CAMC-B082E9/2020\|EPI_ISL_643827\|20 | ···················· | ···················· | ····················· | ···················· | ···················· | ···················· |
| R | 966 | hCoV-19/England/CAMC-B085E0/2020\|EPI_ISL_643830\|20 | ···················· | ···················· | ····················· | ···················· | ···················· | ···················· |
| R | 967 | hCoV-19/England/CAMC-B076FD/2020\|EPI_ISL_643831\|20 | ···················· | ···················· | ····················· | ···················· | ······T············· | ···············A···· |
| R | 968 | hCoV-19/England/CAMC-B08AC3/2020\|EPI_ISL_643832\|20 | ···················· | ···················· | ····················· | ···················· | ···················· | ···················· |
| R | 969 | hCoV-19/England/CAMC-B086CF/2020\|EPI_ISL_643833\|20 | ···················· | ···················· | ····················· | ···················· | ···················· | ···················· |
| R | 970 | hCoV-19/England/CAMC-B07235/2020\|EPI_ISL_643834\|20 | ···················· | ···················· | ····················· | ···················· | ···················· | ···················· |
| R | 971 | hCoV-19/England/CAMC-B0703B/2020\|EPI_ISL_643835\|20 | ···················· | ···················· | ····················· | ···················· | ···················· | ···················· |
| R | 972 | hCoV-19/England/CAMC-B075B4/2020\|EPI_ISL_643837\|20 | ···················· | ···················· | ····················· | ···················· | ···················· | ···················· |
| R | 973 | hCoV-19/England/CAMC-B087EA/2020\|EPI_ISL_643838\|20 | ···················· | ···················· | ····················· | ···················· | ······T············· | ···················· |
| R | 974 | hCoV-19/England/CAMC-B07790/2020\|EPI_ISL_643839\|20 | ···················· | ···················· | ····················· | ···················· | ···················· | ···················· |
| R | 975 | hCoV-19/England/CAMC-B078E8/2020\|EPI_ISL_643840\|20 | ···················· | ···················· | ····················· | ···················· | ···················· | ···················· |
| R | 976 | hCoV-19/England/CAMC-B07086/2020\|EPI_ISL_643842\|20 | ···················· | ···················· | ····················· | ···················· | ···················· | ···················· |
| R | 977 | hCoV-19/England/CAMC-B08331/2020\|EPI_ISL_643854\|20 | ···················· | ···················· | ····················· | ···················· | ···················· | ···················· |
| R | 978 | hCoV-19/England/CAMC-B08735/2020\|EPI_ISL_643855\|20 | ···················· | ···················· | ····················· | ···················· | ···················· | ···················· |
| R | 979 | hCoV-19/England/CAMC-B07CEC/2020\|EPI_ISL_643856\|20 | ···················· | ···················· | ····················· | ···················· | ···················· | ···················· |
| R | 980 | hCoV-19/England/CAMC-B085C2/2020\|EPI_ISL_643857\|20 | ···················· | ···················· | ····················· | ···················· | ···················· | ···················· |
| R | 981 | hCoV-19/England/CAMC-B07F98/2020\|EPI_ISL_643860\|20 | ···················· | ···················· | ····················· | ···················· | ···················· | ···················· |
| R | 982 | hCoV-19/England/CAMC-B077EB/2020\|EPI_ISL_643861\|20 | ···················· | ···················· | ····················· | ···················· | ···················· | ···················· |
| R | 983 | hCoV-19/England/CAMC-B09242/2020\|EPI_ISL_643862\|20 | ···················· | ···················· | ····················· | ···················· | ···················· | ···················· |
| R | 984 | hCoV-19/England/CAMC-B07D8F/2020\|EPI_ISL_643864\|20 | ···················· | ···················· | ····················· | ···················· | ······T············· | ···················· |
| R | 985 | hCoV-19/England/ALDP-B1BD97/2020\|EPI_ISL_646060\|20 | ···················· | ···················· | ····················· | ···················· | ···················· | ···················· |
| R | 986 | hCoV-19/England/ALDP-B1F926/2020\|EPI_ISL_646061\|20 | ···················· | ·A·················· | ····················· | ···················· | ···················· | ···A················ |
| R | 987 | hCoV-19/England/ALDP-B1B25C/2020\|EPI_ISL_646062\|20 | ···················· | ···················· | ····················· | ···················· | ···················· | ···················· |
| R | 988 | hCoV-19/England/ALDP-B1C473/2020\|EPI_ISL_646063\|20 | ···················· | ···················· | ····················· | ···················· | ···················· | ···················· |
| R | 989 | hCoV-19/England/ALDP-B1D612/2020\|EPI_ISL_646064\|20 | ···················· | ···················· | ····················· | ···················· | ···················· | ···················· |
| R | 990 | hCoV-19/England/ALDP-B1DB5F/2020\|EPI_ISL_646066\|20 | ···················· | ···················· | ····················· | ···················· | ···················· | ···················· |
| R | 991 | hCoV-19/England/CAMC-B21E7D/2020\|EPI_ISL_646067\|20 | ···················· | ···················· | ····················· | ···················· | ···················· | ···················· |
| R | 992 | hCoV-19/England/ALDP-B1F971/2020\|EPI_ISL_646068\|20 | ···················· | ···················· | ····················· | ···················· | ···················· | ···················· |
| R | 993 | hCoV-19/England/ALDP-B1FA14/2020\|EPI_ISL_646069\|20 | ···················· | ···················· | ····················· | ···················· | ···················· | ···················· |
| R | 994 | hCoV-19/England/CAMC-B222E9/2020\|EPI_ISL_646070\|20 | ···················· | ·A·················· | ····················· | ···················· | ···················· | ···A················ |
| R | 995 | hCoV-19/England/ALDP-B1A31E/2020\|EPI_ISL_646071\|20 | ···················· | ···················· | ····················· | ···················· | ···················· | ···················· |
| R | 996 | hCoV-19/England/ALDP-B1C4A0/2020\|EPI_ISL_646072\|20 | ···················· | ···················· | ····················· | ···················· | ···················· | ···················· |
| R | 997 | hCoV-19/England/ALDP-B1F838/2020\|EPI_ISL_646073\|20 | ···················· | ···················· | ····················· | ···················· | ···················· | ···················· |
| R | 998 | hCoV-19/England/ALDP-B1B869/2020\|EPI_ISL_646074\|20 | ···················· | ···················· | ····················· | ···················· | ···················· | ···················· |
| R | 999 | hCoV-19/England/ALDP-B1F5B9/2020\|EPI_ISL_646075\|20 | ···················· | ···················· | ····················· | ···················· | ···················· | ···················· |
| R | 1000 | hCoV-19/England/CAMC-B225B3/2020\|EPI_ISL_646076\|20 | ···················· | ···················· | ····················· | ···················· | ···················· | ···················· |
| R | 1001 | hCoV-19/England/ALDP-B1B2C5/2020\|EPI_ISL_646078\|20 | ···················· | ···················· | ····················· | ···················· | ···················· | ···················· |
| R | 1002 | hCoV-19/England/ALDP-B1BD79/2020\|EPI_ISL_646079\|20 | ···················· | ···················· | ····················· | ···················· | ···················· | ···················· |
| R | 1003 | hCoV-19/England/ALDP-B1AD2F/2020\|EPI_ISL_646080\|20 | ···················· | ···················· | ····················· | ···················· | ···················· | ···················· |
| R | 1004 | hCoV-19/England/ALDP-B1AEB3/2020\|EPI_ISL_646081\|20 | ···················· | ···················· | ····················· | ···················· | ···················· | ···················· |
| R | 1005 | hCoV-19/England/ALDP-B1BD6A/2020\|EPI_ISL_646082\|20 | ···················· | ···················· | ····················· | ···················· | ···················· | ···················· |
| R | 1006 | hCoV-19/England/CAMC-B22270/2020\|EPI_ISL_646083\|20 | ···················· | ···················· | ····················· | ···················· | ···················· | ···················· |
| R | 1007 | hCoV-19/England/CAMC-B21BFE/2020\|EPI_ISL_646084\|20 | ···················· | ···················· | ····················· | ···················· | ···················· | ···················· |
| R | 1008 | hCoV-19/England/ALDP-B1AD89/2020\|EPI_ISL_646087\|20 | ···················· | ···················· | ····················· | ···················· | ···················· | ···A················ |
| R | 1009 | hCoV-19/England/ALDP-B1A378/2020\|EPI_ISL_646088\|20 | ···················· | ···················· | ····················· | ···················· | ···················· | ···················· |
| R | 1010 | hCoV-19/England/ALDP-B1ADA7/2020\|EPI_ISL_646090\|20 | ···················· | ···················· | ····················· | ···················· | ···················· | ···················· |
| R | 1011 | hCoV-19/England/CAMC-B2238C/2020\|EPI_ISL_646091\|20 | ···················· | ···················· | ····················· | ···················· | ···················· | ···················· |
| R | 1012 | hCoV-19/England/ALDP-B1F786/2020\|EPI_ISL_646092\|20 | ···················· | ···················· | ····················· | ···················· | ······T············· | ···················· |
| R | 1013 | hCoV-19/England/ALDP-B1A396/2020\|EPI_ISL_646093\|20 | ···················· | ···················· | ····················· | ···················· | ···················· | ···················· |
| R | 1014 | hCoV-19/England/ALDP-B1AC6E/2020\|EPI_ISL_646095\|20 | ···················· | ···················· | ····················· | ···················· | ···················· | ···················· |
| R | 1015 | hCoV-19/England/ALDP-B1BCC7/2020\|EPI_ISL_646097\|20 | ···················· | ···················· | ····················· | ···················· | ···················· | ···················· |
| R | 1016 | hCoV-19/England/ALDP-B1B1C8/2020\|EPI_ISL_646098\|20 | ···················· | ···················· | ····················· | ···················· | ···················· | ···················· |
| R | 1017 | hCoV-19/England/ALDP-B1AF38/2020\|EPI_ISL_646099\|20 | ···················· | ···················· | ····················· | ···················· | ···················· | ···················· |
| R | 1018 | hCoV-19/England/CAMC-B21F89/2020\|EPI_ISL_647390\|20 | ···················· | ···················· | ····················· | ···················· | ···················· | ···················· |
| R | 1019 | hCoV-19/England/CAMC-B22735/2020\|EPI_ISL_647392\|20 | ···················· | ···················· | ····················· | ···················· | ···················· | ···················· |
| R | 1020 | hCoV-19/England/CAMC-B21842/2020\|EPI_ISL_647393\|20 | ···················· | ···················· | ····················· | ···················· | ···················· | ···················· |
| R | 1021 | hCoV-19/England/CAMC-B22CCD/2020\|EPI_ISL_647394\|20 | ···················· | ···················· | ····················· | ···················· | ···················· | ···················· |
| R | 1022 | hCoV-19/England/CAMC-B22FD3/2020\|EPI_ISL_647395\|20 | ···················· | ···················· | ····················· | ···················· | ···················· | ···················· |
| R | 1023 | hCoV-19/England/CAMC-B22F5B/2020\|EPI_ISL_647396\|20 | ···················· | ···················· | ····················· | ···················· | ···················· | ···················· |
| R | 1024 | hCoV-19/England/CAMC-B21A3D/2020\|EPI_ISL_647398\|20 | ···················· | ·A·······A·········· | ····················· | ···················· | ···················· | ···················· |
| R | 1025 | hCoV-19/Australia/VIC15974/2020\|EPI_ISL_641100\|202 | ···················· | ···················· | ····················· | ···················· | ···················· | ···················· |
| R | 1026 | hCoV-19/Australia/VIC12053/2020\|EPI_ISL_641101\|202 | ···················· | ···················· | ····················· | ···················· | ···················· | ···················· |
| R | 1027 | hCoV-19/Australia/VIC12097/2020\|EPI_ISL_641103\|202 | ···················· | ···················· | ····················· | ···················· | ···················· | ···················· |
| R | 1028 | hCoV-19/Australia/VIC16576/2020\|EPI_ISL_641170\|202 | ···················· | ···················· | ····················· | ···················· | ···················· | ···················· |
| R | 1029 | hCoV-19/Australia/VIC16028/2020\|EPI_ISL_641171\|202 | ···················· | ···················· | ····················· | ···················· | ···················· | ···················· |
| R | 1031 | hCoV-19/Australia/VIC12054/2020\|EPI_ISL_641173\|202 | ···················· | ···················· | ····················· | ···················· | ···················· | ················K··· |
| R | 1032 | hCoV-19/Australia/VIC12335/2020\|EPI_ISL_641174\|202 | ···················· | ···················· | ····················· | ···················· | ···················· | ···················· |
| R | 1033 | hCoV-19/Australia/VIC13155/2020\|EPI_ISL_641175\|202 | ···················· | ···················· | ····················· | ···················· | ···················· | ···················· |
| R | 1034 | hCoV-19/Australia/VIC15977/2020\|EPI_ISL_641176\|202 | ···················· | ···················· | ····················· | ···················· | ···················· | ···················· |
| R | 1035 | hCoV-19/Australia/VIC15880/2020\|EPI_ISL_641177\|202 | ···················· | ···················· | ····················· | ···················· | ···················· | ···················· |

| | | | | | | | | |
|---|---|---|---|---|---|---|---|---|
| R | 1036 | hCoV-19/Australia/VIC16121/2020\|EPI_ISL_641178\|202 | ···················· | ···················· | ···················· | ···················· | ···················· | ···················· |
| R | 1037 | hCoV-19/Australia/VIC16042/2020\|EPI_ISL_641179\|202 | ···················· | ···················· | ···················· | ···················· | ···················· | ···················· |
| R | 1038 | hCoV-19/Australia/VIC11643/2020\|EPI_ISL_641181\|202 | ···················· | ···················· | ···················· | ···················· | ···················· | ···················· |
| R | 1040 | hCoV-19/Australia/VIC16315/2020\|EPI_ISL_641185\|202 | ···················· | ···················· | ···················· | ···················· | ···················· | ···················· |
| R | 1042 | hCoV-19/Australia/VIC16306/2020\|EPI_ISL_641187\|202 | ···················· | ···················· | ···················· | ···················· | ···················· | ···················· |
| R | 1044 | hCoV-19/Australia/VIC13194/2020\|EPI_ISL_641190\|202 | ···················· | ···················· | ···················· | ···················· | ······R············· | ···················· |
| R | 1045 | hCoV-19/Australia/VIC12768/2020\|EPI_ISL_641192\|202 | ···················· | ···················· | ···················· | ···················· | ···················· | ···················· |
| R | 1046 | hCoV-19/Australia/VIC16242/2020\|EPI_ISL_641193\|202 | ···················· | ···················· | ···················· | ···················· | ···················· | ···················· |
| R | 1047 | hCoV-19/Australia/VIC16520/2020\|EPI_ISL_641194\|202 | ···················· | ···················· | ···················· | ···················· | ···················· | ···················· |
| R | 1048 | hCoV-19/Australia/VIC12811/2020\|EPI_ISL_641195\|202 | ···················· | ···················· | ···················· | ···················· | ···················· | ···················· |
| R | 1049 | hCoV-19/Australia/VIC11730/2020\|EPI_ISL_641196\|202 | ···················· | ···················· | ···················· | ···················· | ···················· | ···················· |
| R | 1051 | hCoV-19/Australia/VIC12624/2020\|EPI_ISL_641198\|202 | ···················· | ···················· | ···················· | ···················· | ···················· | ···················· |
| R | 1052 | hCoV-19/Australia/VIC12055/2020\|EPI_ISL_641199\|202 | ···················· | ···················· | ···················· | ···················· | ···················· | ···················· |
| R | 1053 | hCoV-19/England/QEUH-B130C3/2020\|EPI_ISL_642401\|20 | ···················· | ···················· | ···················· | ···················· | ···················· | ···················· |
| R | 1054 | hCoV-19/England/QEUH-B11C94/2020\|EPI_ISL_642402\|20 | ···················· | ···················· | ···················· | ···················· | ···················· | ···················· |
| R | 1055 | hCoV-19/England/QEUH-B10EF9/2020\|EPI_ISL_642403\|20 | ···················· | ···················· | ···················· | ···················· | ···················· | ···················· |
| R | 1056 | hCoV-19/England/QEUH-B11DFB/2020\|EPI_ISL_642404\|20 | ···················· | ···················· | ···················· | ···················· | ···················· | ···················· |
| R | 1057 | hCoV-19/England/QEUH-B12950/2020\|EPI_ISL_642405\|20 | ···················· | ···················· | ···················· | ···················· | ···················· | ···················· |
| R | 1058 | hCoV-19/England/QEUH-B13676/2020\|EPI_ISL_642406\|20 | ···················· | ···················· | ···················· | ···················· | ···················· | ···················· |
| R | 1059 | hCoV-19/England/QEUH-B10F50/2020\|EPI_ISL_642407\|20 | ···················· | ···················· | ···················· | ···················· | ···················· | ···················· |
| R | 1060 | hCoV-19/England/QEUH-B11924/2020\|EPI_ISL_642409\|20 | ···················· | ···················· | ···················· | ···················· | ···················· | ···················· |
| R | 1061 | hCoV-19/England/QEUH-B11D73/2020\|EPI_ISL_642410\|20 | ···················· | ···················· | ···················· | ···················· | ···················· | ···················· |
| R | 1062 | hCoV-19/England/QEUH-B116E1/2020\|EPI_ISL_642412\|20 | ···················· | ·A·················· | ···················· | ···················· | ···················· | ···················· |
| R | 1063 | hCoV-19/England/QEUH-B11906/2020\|EPI_ISL_642423\|20 | ···················· | ···················· | ···················· | ···················· | ···················· | ···················· |
| R | 1064 | hCoV-19/England/QEUH-B11599/2020\|EPI_ISL_642424\|20 | ···················· | ···················· | ···················· | ···················· | ···················· | ···················· |
| R | 1065 | hCoV-19/England/QEUH-B11423/2020\|EPI_ISL_642425\|20 | ···················· | ·A·················· | ···················· | ···················· | ···················· | ···················· |
| R | 1066 | hCoV-19/England/QEUH-B114C9/2020\|EPI_ISL_642426\|20 | ···················· | ···················· | ···················· | ···················· | ···················· | ···················· |
| R | 1067 | hCoV-19/England/QEUH-B114BA/2020\|EPI_ISL_642427\|20 | ···················· | ···················· | ···················· | ···················· | ···················· | ···················· |
| R | 1068 | hCoV-19/England/QEUH-B13157/2020\|EPI_ISL_642428\|20 | ···················· | ···················· | ···················· | ···················· | ···················· | ···················· |
| R | 1069 | hCoV-19/England/QEUH-B116B4/2020\|EPI_ISL_642429\|20 | ···················· | ···················· | ···················· | ···················· | ···················· | ···················· |
| R | 1070 | hCoV-19/England/QEUH-B126D1/2020\|EPI_ISL_642430\|20 | ···················· | ···················· | ···················· | ···················· | ···················· | ···················· |
| R | 1071 | hCoV-19/England/QEUH-B11854/2020\|EPI_ISL_642431\|20 | ···················· | ···················· | ···················· | ···················· | ···················· | ···················· |
| R | 1072 | hCoV-19/England/QEUH-B105A9/2020\|EPI_ISL_642432\|20 | ···················· | ···················· | ···················· | ···················· | ···················· | ···················· |
| R | 1073 | hCoV-19/England/QEUH-B10D56/2020\|EPI_ISL_642433\|20 | ···················· | ···················· | ···················· | ···················· | ···················· | ···················· |
| R | 1074 | hCoV-19/England/QEUH-B10E80/2020\|EPI_ISL_642434\|20 | ···················· | ···················· | ···················· | ···················· | ···················· | ···················· |
| R | 1075 | hCoV-19/England/CAMC-B08C63/2020\|EPI_ISL_643700\|20 | ···················· | ···················· | ···················· | ···················· | ···················· | ···················· |
| R | 1076 | hCoV-19/England/CAMC-B08A3C/2020\|EPI_ISL_643701\|20 | ···················· | ···················· | ···················· | ···················· | ···················· | ···················· |
| R | 1077 | hCoV-19/England/CAMC-B087BD/2020\|EPI_ISL_643702\|20 | ···················· | ···················· | ···················· | ···················· | ···················· | ···················· |
| R | 1078 | hCoV-19/England/CAMC-B072CC/2020\|EPI_ISL_643703\|20 | ···················· | ···················· | ···················· | ···················· | ···················· | ···················· |
| R | 1079 | hCoV-19/England/CAMC-B08B75/2020\|EPI_ISL_643704\|20 | ···················· | ···················· | ···················· | ···················· | ···················· | ···················· |
| R | 1080 | hCoV-19/England/CAMC-B08207/2020\|EPI_ISL_643705\|20 | ···················· | ···················· | ···················· | ···················· | ···················· | ···················· |
| R | 1081 | hCoV-19/England/CAMC-B08191/2020\|EPI_ISL_643706\|20 | ···················· | ···················· | ···················· | ···················· | ······T············· | ···················· |
| R | 1082 | hCoV-19/England/CAMC-B071ED/2020\|EPI_ISL_643707\|20 | ···················· | ···················· | ···················· | ···················· | ···················· | ···················· |
| R | 1083 | hCoV-19/England/CAMC-B07F89/2020\|EPI_ISL_643708\|20 | ···················· | ···················· | ···················· | ···················· | ···················· | ···················· |
| R | 1084 | hCoV-19/England/CAMC-B0798B/2020\|EPI_ISL_643709\|20 | ···················· | ···················· | ···················· | ···················· | ···················· | ···················· |
| R | 1085 | hCoV-19/England/CAMC-B07D70/2020\|EPI_ISL_643711\|20 | ···················· | ···················· | ···················· | ···················· | ···················· | ···················· |
| R | 1086 | hCoV-19/England/CAMC-B0704A/2020\|EPI_ISL_643712\|20 | ···················· | ···················· | ···················· | ···················· | ···················· | ···················· |
| R | 1087 | hCoV-19/England/CAMC-B092CA/2020\|EPI_ISL_643713\|20 | ···················· | ···················· | ···················· | ···················· | ···················· | ···················· |
| R | 1088 | hCoV-19/England/CAMC-B070FF/2020\|EPI_ISL_643714\|20 | ···················· | ···················· | ···················· | ···················· | ···················· | ···················· |
| R | 1089 | hCoV-19/England/CAMC-B07F3E/2020\|EPI_ISL_643715\|20 | ···················· | ···················· | ···················· | ···················· | ···················· | ···················· |
| R | 1090 | hCoV-19/England/CAMC-B07420/2020\|EPI_ISL_643716\|20 | ···················· | ···················· | ···················· | ···················· | ···················· | ···················· |
| R | 1091 | hCoV-19/England/CAMC-B0751E/2020\|EPI_ISL_643717\|20 | ···················· | ···················· | ···················· | ···················· | ···················· | ···················· |
| R | 1092 | hCoV-19/England/CAMC-B0860B/2020\|EPI_ISL_643718\|20 | ···················· | ···················· | ···················· | ···················· | ···················· | ···················· |
| R | 1093 | hCoV-19/England/CAMC-B07323/2020\|EPI_ISL_643719\|20 | ···················· | ···················· | ···················· | ···················· | ···················· | ···················· |
| R | 1094 | hCoV-19/England/CAMC-B086B0/2020\|EPI_ISL_643720\|20 | ···················· | ···················· | ···················· | ···················· | ···················· | ···················· |
| R | 1095 | hCoV-19/England/CAMC-B081EC/2020\|EPI_ISL_643721\|20 | ···················· | ···················· | ···················· | ···················· | ···················· | ···················· |
| R | 1096 | hCoV-19/England/CAMC-B092F7/2020\|EPI_ISL_643722\|20 | ···················· | ···················· | ···················· | ···················· | ···················· | ···················· |
| R | 1097 | hCoV-19/England/CAMC-B079C7/2020\|EPI_ISL_643723\|20 | ···················· | ···················· | ···················· | ···················· | ···················· | ···················· |
| R | 1098 | hCoV-19/England/CAMC-B0796D/2020\|EPI_ISL_643724\|20 | ···················· | ·A·················· | ···················· | ···················· | ···················· | ···A················ |
| R | 1099 | hCoV-19/England/CAMC-B0794F/2020\|EPI_ISL_643726\|20 | ···················· | ···················· | ···················· | ···················· | ···················· | ···················· |
| R | 1100 | hCoV-19/England/CAMC-B077DC/2020\|EPI_ISL_643727\|20 | ···················· | ···················· | ···················· | ···················· | ···················· | ···················· |
| R | 1101 | hCoV-19/England/CAMC-B072DB/2020\|EPI_ISL_643728\|20 | ···················· | ···················· | ···················· | ···················· | ···················· | ···················· |
| R | 1102 | hCoV-19/England/CAMC-B07D07/2020\|EPI_ISL_643729\|20 | ···················· | ···················· | ···················· | ···················· | ···················· | ···················· |
| R | 1103 | hCoV-19/England/CAMC-B07763/2020\|EPI_ISL_643730\|20 | ···················· | ···················· | ···················· | ···················· | ···················· | ···················· |
| R | 1104 | hCoV-19/England/CAMC-B085FF/2020\|EPI_ISL_643731\|20 | ···················· | ···················· | ···················· | ···················· | ···················· | ···················· |
| R | 1105 | hCoV-19/England/CAMC-B07CFB/2020\|EPI_ISL_643732\|20 | ···················· | ···················· | ···················· | ···················· | ···················· | ···················· |
| R | 1106 | hCoV-19/England/CAMC-B092AC/2020\|EPI_ISL_643733\|20 | ···················· | ···················· | ···················· | ···················· | ···················· | ···················· |
| R | 1107 | hCoV-19/England/CAMC-B0889C/2020\|EPI_ISL_643734\|20 | ···················· | ·A·················· | ···················· | ···················· | ···················· | ···A················ |
| R | 1108 | hCoV-19/England/CAMC-B07C55/2020\|EPI_ISL_643735\|20 | ···················· | ···················· | ···················· | ···················· | ···················· | ···················· |
| R | 1109 | hCoV-19/England/CAMC-B075A5/2020\|EPI_ISL_643736\|20 | ···················· | ···················· | ···················· | ···················· | ···················· | ···················· |
| R | 1110 | hCoV-19/England/CAMC-B08683/2020\|EPI_ISL_643737\|20 | ···················· | ···················· | ···················· | ···················· | ···················· | ···················· |
| R | 1111 | hCoV-19/England/CAMC-B07912/2020\|EPI_ISL_643738\|20 | ···················· | ···················· | ···················· | ···················· | ···················· | ···················· |
| R | 1112 | hCoV-19/England/CAMC-B08A2D/2020\|EPI_ISL_643739\|20 | ···················· | ···················· | ···················· | ···················· | ···················· | ···················· |
| R | 1113 | hCoV-19/England/CAMC-B082F8/2020\|EPI_ISL_643740\|20 | ···················· | ···················· | ···················· | ···················· | ···················· | ···················· |
| R | 1114 | hCoV-19/England/CAMC-B08A96/2020\|EPI_ISL_643742\|20 | ···················· | ·A·················· | ···················· | ···················· | ···················· | ···A················ |
| R | 1115 | hCoV-19/England/CAMC-B071B0/2020\|EPI_ISL_643743\|20 | ···················· | ···················· | ···················· | ···················· | ···················· | ···················· |
| R | 1116 | hCoV-19/England/CAMC-B086FC/2020\|EPI_ISL_643755\|20 | ···················· | ···················· | ···················· | ···················· | ···················· | ···················· |
| R | 1117 | hCoV-19/England/CAMC-B0736F/2020\|EPI_ISL_643756\|20 | ···················· | ···················· | ···················· | ···················· | ···················· | ···················· |
| R | 1118 | hCoV-19/England/CAMC-B08322/2020\|EPI_ISL_643757\|20 | ···················· | ···················· | ···················· | ···················· | ···················· | ···················· |
| R | 1119 | hCoV-19/England/CAMC-B0745D/2020\|EPI_ISL_643758\|20 | ···················· | ···················· | ···················· | ···················· | ···················· | ···················· |

R 1120 hCoV-19/England/CAMC-B08155/2020|EPI_ISL_643760|20 ···················· ···················· ····················· ···················· ···················· ····················
R 1121 hCoV-19/England/CAMC-B08A4B/2020|EPI_ISL_643761|20 ···················· ·A·················· ····················· ···················· ···················· ··A·················
R 1122 hCoV-19/England/CAMC-B08A5A/2020|EPI_ISL_643762|20 ···················· ···················· ····················· ···················· ···················· ····················
R 1123 hCoV-19/England/CAMC-B07CBF/2020|EPI_ISL_643763|20 ···················· ···················· ····················· ···················· ·····T·············· ····················
R 1124 hCoV-19/England/CAMC-B0739C/2020|EPI_ISL_643764|20 ···················· ···················· ····················· ···················· ···················· ····················
R 1125 hCoV-19/England/CAMC-B07A1F/2020|EPI_ISL_643765|20 ···················· ···················· ····················· ···················· ···················· ····················
R 1126 hCoV-19/England/CAMC-B21411/2020|EPI_ISL_647200|20 ···················· ···················· ····················· ···················· ···················· ····················
R 1127 hCoV-19/England/CAMC-B216EE/2020|EPI_ISL_647201|20 ···················· ···················· ····················· ···················· ···················· ····················
R 1128 hCoV-19/England/CAMC-B226A1/2020|EPI_ISL_647202|20 ···················· ···················· ····················· ···················· ···················· ····················
R 1129 hCoV-19/England/CAMC-B2279F/2020|EPI_ISL_647203|20 ···················· ···················· ····················· ···················· ···················· ····················
R 1130 hCoV-19/England/CAMC-B22489/2020|EPI_ISL_647204|20 ···················· ···················· ····················· ···················· ···················· ····················
R 1131 hCoV-19/England/CAMC-B2187F/2020|EPI_ISL_647205|20 ···················· ···················· ····················· ···················· ···················· ····················
R 1132 hCoV-19/England/CAMC-B22F2E/2020|EPI_ISL_647206|20 ···················· ···················· ····················· ···················· ···················· ····················
R 1133 hCoV-19/England/CAMC-B22EC7/2020|EPI_ISL_647208|20 ···················· ···················· ····················· ···················· ···················· ····················
R 1134 hCoV-19/England/CAMC-B227CC/2020|EPI_ISL_647209|20 ···················· ···················· ····················· ···················· ···················· ····················
R 1135 hCoV-19/England/CAMC-B2151E/2020|EPI_ISL_647210|20 ···················· ···················· ····················· ···················· ···················· ····················
R 1136 hCoV-19/England/CAMC-B22F1F/2020|EPI_ISL_647212|20 ···················· ···················· ····················· ···················· ···················· ····················
R 1137 hCoV-19/England/CAMC-B2145D/2020|EPI_ISL_647213|20 ···················· ···················· ····················· ···················· ···················· ····················
R 1138 hCoV-19/England/CAMC-B22B93/2020|EPI_ISL_647215|20 ···················· ···················· ····················· ···················· ···················· ····················
R 1139 hCoV-19/England/CAMC-B21AB5/2020|EPI_ISL_647216|20 ···················· ···················· ····················· ···················· ···················· ····················
R 1140 hCoV-19/England/CAMC-B214B7/2020|EPI_ISL_647218|20 ···················· ···················· ····················· ···················· ···················· ····················
R 1141 hCoV-19/England/CAMC-B22A1E/2020|EPI_ISL_647221|20 ···················· ···················· ····················· ···················· ···················· ····················
R 1142 hCoV-19/England/CAMC-B21F6B/2020|EPI_ISL_647222|20 ···················· ···················· ····················· ···················· ···················· ····················
R 1143 hCoV-19/England/CAMC-B22EB8/2020|EPI_ISL_647224|20 ···················· ···················· ····················· ···················· ·····T·············· ····················
R 1144 hCoV-19/England/CAMC-B22A5A/2020|EPI_ISL_647225|20 ···················· ···················· ····················· ···················· ···················· ····················
R 1145 hCoV-19/England/CAMC-B21806/2020|EPI_ISL_647227|20 ···················· ·A·················· ····················· ···················· ···················· ··A·················
R 1146 hCoV-19/England/CAMC-B217EB/2020|EPI_ISL_647228|20 ···················· ···················· ····················· ···················· ···················· ····················
R 1147 hCoV-19/England/CAMC-B21DAD/2020|EPI_ISL_647230|20 ···················· ·A·················· ····················· ···················· ···················· ··A·················
R 1148 hCoV-19/England/CAMC-B217FA/2020|EPI_ISL_647231|20 ···················· ···················· ····················· ···················· ···T················ ····················
R 1149 hCoV-19/England/CAMC-B22D8E/2020|EPI_ISL_647232|20 ···················· ···················· ····················· ···················· ···················· ····················
R 1150 hCoV-19/England/CAMC-B2252C/2020|EPI_ISL_647233|20 ···················· ···················· ····················· ···················· ···················· ····················
R 1151 hCoV-19/England/CAMC-B216C0/2020|EPI_ISL_647234|20 ···················· ···················· ····················· ···················· ···················· ····················
R 1152 hCoV-19/England/CAMC-B221EC/2020|EPI_ISL_647235|20 ···················· ···················· ····················· ···················· ···················· ····················
R 1153 hCoV-19/England/CAMC-B22D15/2020|EPI_ISL_647236|20 ···················· ···················· ····················· ···················· ···················· ····················
R 1154 hCoV-19/England/CAMC-B229D5/2020|EPI_ISL_647238|20 ···················· ···················· ····················· ···················· ···················· ····················
R 1155 hCoV-19/England/CAMC-B2250E/2020|EPI_ISL_647239|20 ···················· ···················· ····················· ···················· ···················· ····················
R 1156 hCoV-19/England/CAMC-B21B1C/2020|EPI_ISL_647240|20 ···················· ···················· ····················· ···················· ···················· ····················
R 1157 hCoV-19/England/CAMC-B21077/2020|EPI_ISL_647241|20 ···················· ···················· ····················· ···················· ···················· ····················
R 1158 hCoV-19/England/CAMC-B22762/2020|EPI_ISL_647242|20 ···················· ···················· ····················· ···················· ···················· ····················
R 1159 hCoV-19/England/CAMC-B21C28/2020|EPI_ISL_647243|20 ···················· ···················· ····················· ···················· ···················· ····················
R 1160 hCoV-19/England/CAMC-B227BD/2020|EPI_ISL_647244|20 ···················· ···················· ····················· ···················· ···················· ····················
R 1161 hCoV-19/England/CAMC-B22F00/2020|EPI_ISL_647245|20 ···················· ···················· ····················· ···················· ···················· ····················
R 1162 hCoV-19/England/CAMC-B22692/2020|EPI_ISL_647246|20 ···················· ···················· ····················· ···················· ···················· ····················
R 1163 hCoV-19/England/CAMC-B21B58/2020|EPI_ISL_647247|20 ···················· ···················· ····················· ···················· ···················· ····················
R 1164 hCoV-19/England/CAMC-B21341/2020|EPI_ISL_647249|20 ···················· ···················· ····················· ···················· ···················· ····················
R 1165 hCoV-19/England/CAMC-B218F7/2020|EPI_ISL_647251|20 ···················· ···················· ····················· ···················· ···················· ····················
R 1166 hCoV-19/England/CAMC-B21A1F/2020|EPI_ISL_647252|20 ···················· ···················· ····················· ···················· ···················· ····················
R 1167 hCoV-19/England/CAMC-B2297B/2020|EPI_ISL_647253|20 ···················· ···················· ····················· ···················· ···················· ····················
R 1168 hCoV-19/England/CAMC-B228AB/2020|EPI_ISL_647254|20 ···················· ···················· ····················· ···················· ···················· ····················
R 1169 hCoV-19/England/CAMC-B21CA0/2020|EPI_ISL_647256|20 ···················· ···················· ····················· ···················· ···T················ ····················
R 1170 hCoV-19/England/CAMC-B22E8B/2020|EPI_ISL_647257|20 ···················· ···················· ····················· ···················· ···················· ····················
R 1171 hCoV-19/England/CAMC-B21B76/2020|EPI_ISL_647259|20 ···················· ···················· ····················· ···················· ···················· ····················
R 1172 hCoV-19/England/CAMC-B21DCB/2020|EPI_ISL_647260|20 ···················· ···················· ····················· ···················· ···················· ····················
R 1173 hCoV-19/England/CAMC-B2150F/2020|EPI_ISL_647262|20 ···················· ···················· ····················· ···················· ···················· ····················
R 1174 hCoV-19/England/CAMC-B21AA6/2020|EPI_ISL_647263|20 ···················· ···················· ····················· ···················· ···················· ····················
R 1175 hCoV-19/England/CAMC-B22F88/2020|EPI_ISL_647265|20 ···················· ···················· ····················· ···················· ···················· ····················
R 1176 hCoV-19/England/CAMC-B218BB/2020|EPI_ISL_647266|20 ···················· ···················· ····················· ···················· ···················· ····················
R 1177 hCoV-19/England/CAMC-B22DAC/2020|EPI_ISL_647267|20 ···················· ···················· ····················· ···················· ···················· ····················
R 1178 hCoV-19/England/CAMC-B224A7/2020|EPI_ISL_647268|20 ···················· ···················· ····················· ···················· ·····T·············· ····················
R 1179 hCoV-19/England/CAMC-B21F7A/2020|EPI_ISL_647276|20 ···················· ···················· ····················· ···················· ···················· ····················
R 1180 hCoV-19/England/CAMC-B21CDD/2020|EPI_ISL_647290|20 ···················· ···················· ····················· ···················· ···T················ ····················
R 1181 hCoV-19/England/CAMC-B227EA/2020|EPI_ISL_647291|20 ···················· ···················· ····················· ···················· ···················· ····················
R 1182 hCoV-19/England/CAMC-B216DF/2020|EPI_ISL_647292|20 ···················· ···················· ····················· ···················· ···················· ····················
R 1183 hCoV-19/England/CAMC-B22999/2020|EPI_ISL_647293|20 ···················· ···················· ····················· ···················· ···················· ····················
R 1184 hCoV-19/England/CAMC-B21AC4/2020|EPI_ISL_647294|20 ···················· ···················· ····················· ···················· ···················· ····················
R 1185 hCoV-19/England/CAMC-B21AF1/2020|EPI_ISL_647296|20 ···················· ···················· ····················· ···················· ···················· ····················
R 1186 hCoV-19/England/CAMC-B22AB4/2020|EPI_ISL_647297|20 ···················· ···················· ····················· ···················· ···················· ····················
R 1187 hCoV-19/England/CAMC-B226DE/2020|EPI_ISL_647298|20 ···················· ···················· ····················· ···················· ···················· ····················
R 1188 hCoV-19/England/CAMC-B22DBB/2020|EPI_ISL_647299|20 ···················· ···················· ····················· ···················· ···················· ····················
R 1189 hCoV-19/England/CAMC-B22164/2020|EPI_ISL_647302|20 ···················· ···················· ····················· ···················· ···················· ····················
R 1190 hCoV-19/England/CAMC-B232D9/2020|EPI_ISL_647304|20 ···················· ···················· ····················· ···················· ···················· ····················
R 1191 hCoV-19/England/CAMC-B21350/2020|EPI_ISL_647305|20 ···················· ···················· ····················· ···················· ···················· ····················
R 1192 hCoV-19/England/CAMC-B22E9A/2020|EPI_ISL_647306|20 ···················· ···················· ····················· ···················· ···················· ····················
R 1193 hCoV-19/England/CAMC-B22744/2020|EPI_ISL_647307|20 ···················· ···················· ····················· ···················· ···················· ····················
R 1194 hCoV-19/USA/CA-CZB-12862/2020|EPI_ISL_648518|2020- ···················· ···················· ····················· ···················· ···················· ····················
R 1195 hCoV-19/USA/CA-CZB-12864/2020|EPI_ISL_648519|2020- ···················· ···················· ····················· ···················· ···················· ····················
R 1196 hCoV-19/USA/CA-CZB-12865/2020|EPI_ISL_648520|2020- ···················· ·A·················· ····················· ···················· ···················· ····················
R 1197 hCoV-19/USA/CA-CZB-12867/2020|EPI_ISL_648522|2020- ···················· ···················· ····················· ···················· ···················· ····················
R 1198 hCoV-19/USA/CA-CZB-12868/2020|EPI_ISL_648523|2020- ···················· ···················· ····················· ···················· ···················· ····················
R 1199 hCoV-19/USA/CA-CZB-12869/2020|EPI_ISL_648524|2020- ···················· ···················· ····················· ···················· ···················· ····················

| | | | | | | | | |
|---|---|---|---|---|---|---|---|---|
| R | 1200 | hCoV-19/USA/CA-CZB-12870/2020\|EPI_ISL_648525\|2020- | ···················· | ···················· | ···················· | ···················· | ···················· | ···················· |
| R | 1201 | hCoV-19/USA/CA-CZB-12871/2020\|EPI_ISL_648526\|2020- | ···················· | ···················· | ···················· | ···················· | ···················· | ···················· |
| R | 1202 | hCoV-19/USA/CA-CZB-12872/2020\|EPI_ISL_648527\|2020- | ···················· | ···················· | ···················· | ···················· | ···················· | ···················· |
| R | 1203 | hCoV-19/USA/CA-CZB-12874/2020\|EPI_ISL_648529\|2020- | ···················· | ···················· | ···················· | ···················· | ···················· | ···················· |
| R | 1204 | hCoV-19/USA/CA-CZB-12875/2020\|EPI_ISL_648530\|2020- | ···················· | ···················· | ···················· | ···················· | ···················· | ···················· |
| R | 1205 | hCoV-19/USA/CA-CZB-12370/2020\|EPI_ISL_648531\|2020- | ···················· | ···················· | ···················· | ···················· | ···················· | ···················· |
| R | 1206 | hCoV-19/USA/CA-CZB-12371/2020\|EPI_ISL_648532\|2020- | ···················· | ···················· | ···················· | ···················· | ···················· | ···················· |
| R | 1207 | hCoV-19/USA/CA-CZB-12372/2020\|EPI_ISL_648533\|2020- | ···················· | ···················· | ···················· | ···················· | ···················· | ···················· |
| R | 1208 | hCoV-19/USA/CA-CZB-12373/2020\|EPI_ISL_648534\|2020- | ···················· | ···················· | ···················· | ···················· | ···················· | ···················· |
| R | 1209 | hCoV-19/USA/CA-CZB-12374/2020\|EPI_ISL_648535\|2020- | ···················· | ···················· | ···················· | ···················· | ···················· | ···················· |
| R | 1210 | hCoV-19/USA/CA-CZB-12375/2020\|EPI_ISL_648536\|2020- | ···················· | ···················· | ···················· | ···················· | ···················· | ···················· |
| R | 1211 | hCoV-19/USA/CA-CZB-12376/2020\|EPI_ISL_648537\|2020- | ···················· | ···················· | ···················· | ···················· | ···················· | ···················· |
| R | 1212 | hCoV-19/USA/CA-CZB-12377/2020\|EPI_ISL_648538\|2020- | ···················· | ···················· | ···················· | ···················· | ···················· | ···················· |
| R | 1213 | hCoV-19/USA/CA-CZB-12378/2020\|EPI_ISL_648539\|2020- | ···················· | ·A·················· | ···················· | ···················· | ···················· | ···················· |
| R | 1214 | hCoV-19/USA/CA-CZB-12379/2020\|EPI_ISL_648540\|2020- | ···················· | ···················· | ···················· | ···················· | ···················· | ···················· |
| R | 1215 | hCoV-19/USA/CA-CZB-12380/2020\|EPI_ISL_648541\|2020- | ···················· | ·A·················· | ···················· | ···················· | ···················· | ···················· |
| R | 1216 | hCoV-19/USA/CA-CZB-2811/2020\|EPI_ISL_648542\|2020-0 | ···················· | ·A·················· | ···················· | ···················· | ···················· | ···················· |
| R | 1217 | hCoV-19/USA/CA-CZB-10364/2020\|EPI_ISL_648545\|2020- | ···················· | ·A·················· | ···················· | ···················· | ···················· | ···················· |
| R | 1218 | hCoV-19/USA/CA-CZB-10362/2020\|EPI_ISL_648546\|2020- | ···················· | ·A·················· | ···················· | ···················· | ···················· | ···················· |
| R | 1219 | hCoV-19/USA/CA-CZB-10361/2020\|EPI_ISL_648547\|2020- | ···················· | ·A·················· | ···················· | ···················· | ···················· | ···················· |
| R | 1220 | hCoV-19/USA/CA-CZB-10360/2020\|EPI_ISL_648548\|2020- | ···················· | ·A·················· | ···················· | ···················· | ···················· | ···················· |
| R | 1221 | hCoV-19/USA/CA-CZB-10359/2020\|EPI_ISL_648549\|2020- | ···················· | ···A················ | ···················· | ···················· | ···················· | ···················· |
| R | 1222 | hCoV-19/USA/CA-CZB-10622/2020\|EPI_ISL_648550\|2020- | ···················· | ···················· | ···················· | ···················· | ···················· | ···················· |
| R | 1223 | hCoV-19/USA/CA-CZB-10621/2020\|EPI_ISL_648551\|2020- | ···················· | ·A·················· | ···················· | ···················· | ···················· | ···················· |
| R | 1224 | hCoV-19/USA/CA-CZB-10620/2020\|EPI_ISL_648552\|2020- | ···················· | ·A·················· | ···················· | ···················· | ···················· | ···················· |
| R | 1225 | hCoV-19/USA/CA-CZB-10619/2020\|EPI_ISL_648553\|2020- | ···················· | ·A·················· | ···················· | ···················· | ···················· | ···················· |
| R | 1226 | hCoV-19/USA/CA-CZB-10617/2020\|EPI_ISL_648554\|2020- | ···················· | ···················· | ···················· | ···················· | ···················· | ···················· |
| R | 1227 | hCoV-19/USA/UT-UPHL-201131133/2020\|EPI_ISL_648556\| | ···················· | ···················· | ···················· | ···················· | ···················· | ···················· |
| R | 1228 | hCoV-19/USA/UT-UPHL-201113704/2020\|EPI_ISL_648558\| | ···················· | ···················· | ···················· | ···················· | ···················· | ···················· |
| R | 1229 | hCoV-19/USA/UT-UPHL-201118773/2020\|EPI_ISL_648561\| | ···················· | ···················· | ···················· | ···················· | ···················· | ···················· |
| R | 1230 | hCoV-19/USA/UT-UPHL-201121351/2020\|EPI_ISL_648568\| | ···················· | ···················· | ···················· | ···················· | ···················· | ···················· |
| R | 1231 | hCoV-19/USA/UT-UPHL-201126219/2020\|EPI_ISL_648569\| | ···················· | ···················· | ···················· | ···················· | ···················· | ···················· |
| R | 1232 | hCoV-19/USA/UT-UPHL-201108622/2020\|EPI_ISL_648571\| | ···················· | ···················· | ···················· | ···················· | ···················· | ···················· |
| R | 1233 | hCoV-19/USA/UT-UPHL-201127728/2020\|EPI_ISL_648572\| | ···················· | ···················· | ···················· | ···················· | ···················· | ···················· |
| R | 1234 | hCoV-19/USA/UT-UPHL-201129708/2020\|EPI_ISL_648573\| | ···················· | ···················· | ···················· | ···················· | ···················· | ···················· |
| R | 1235 | hCoV-19/USA/UT-UPHL-201117199/2020\|EPI_ISL_648574\| | ···················· | ···················· | ···················· | ···················· | ···················· | ···················· |
| R | 1236 | hCoV-19/USA/UT-UPHL-201107239/2020\|EPI_ISL_648575\| | ···················· | ···················· | ···················· | ···················· | ···················· | ···················· |
| R | 1237 | hCoV-19/USA/UT-UPHL-201113629/2020\|EPI_ISL_648576\| | ···················· | ···················· | ···················· | ···················· | ···················· | ···················· |
| R | 1238 | hCoV-19/USA/UT-UPHL-201112041/2020\|EPI_ISL_648578\| | ···················· | ···················· | ···················· | ···················· | ···················· | ···················· |
| R | 1239 | hCoV-19/USA/UT-UPHL-201117069/2020\|EPI_ISL_648579\| | ···················· | ·A·················· | ···················· | ···················· | ···················· | ···················· |
| R | 1240 | hCoV-19/USA/UT-UPHL-201103661/2020\|EPI_ISL_648580\| | ···················· | ···················· | ···················· | ···················· | ···················· | ···················· |
| R | 1241 | hCoV-19/USA/UT-UPHL-201128010/2020\|EPI_ISL_648582\| | ···················· | ···················· | ···················· | ···················· | ···················· | ···················· |
| R | 1242 | hCoV-19/USA/UT-UPHL-201108464/2020\|EPI_ISL_648585\| | ···················· | ·A·················· | ···················· | ···················· | ···················· | ···················· |
| R | 1243 | hCoV-19/USA/UT-UPHL-201108102/2020\|EPI_ISL_648586\| | ···················· | ···················· | ···················· | ···················· | ···················· | ···················· |
| R | 1244 | hCoV-19/USA/UT-UPHL-201116393/2020\|EPI_ISL_648588\| | ···················· | ···················· | ···················· | ···················· | ···················· | ···················· |
| R | 1245 | hCoV-19/USA/UT-UPHL-201131465/2020\|EPI_ISL_648590\| | ···················· | ···················· | ···················· | ···················· | ···················· | ···················· |
| R | 1246 | hCoV-19/USA/UT-UPHL-201104262/2020\|EPI_ISL_648591\| | ···················· | ···················· | ···················· | ···················· | ···················· | ···················· |
| R | 1247 | hCoV-19/USA/UT-UPHL-201116223/2020\|EPI_ISL_648592\| | ···················· | ···················· | ···················· | ···················· | ···················· | ···················· |
| R | 1248 | hCoV-19/Mexico/NLE-UANL-126/2020\|EPI_ISL_648595\|20 | ···················· | ···················· | ···················· | ···················· | ···················· | ···················· |
| R | 1249 | hCoV-19/Mexico/NLE-UANL-2098/2020\|EPI_ISL_648602\|2 | ···················· | ···················· | ···················· | ···················· | ···················· | ···················· |
| R | 1250 | hCoV-19/Mexico/NLE-UANL-2088/2020\|EPI_ISL_648603\|2 | ···················· | ···················· | ···················· | ···················· | ···················· | ···················· |
| R | 1251 | hCoV-19/Mexico/NLE-UANL-4479/2020\|EPI_ISL_648607\|2 | ···················· | ···················· | ···················· | ···················· | ···················· | ···················· |
| R | 1252 | hCoV-19/Singapore/1194/2020\|EPI_ISL_648610\|2020-04 | ···················· | ···················· | ···················· | ···················· | ···················· | ···················· |
| R | 1253 | hCoV-19/Singapore/1195/2020\|EPI_ISL_648611\|2020-04 | ···················· | ···················· | ···················· | ···················· | ···················· | ···················· |
| R | 1254 | hCoV-19/Singapore/1196/2020\|EPI_ISL_648612\|2020-04 | ···················· | ···················· | ···················· | ···················· | ···················· | ···················· |
| R | 1255 | hCoV-19/Singapore/1197/2020\|EPI_ISL_648613\|2020-04 | ···················· | ···················· | ···················· | ···················· | ···················· | ···················· |
| R | 1256 | hCoV-19/Singapore/1198/2020\|EPI_ISL_648614\|2020-04 | ···················· | ···················· | ···················· | ···················· | ···················· | ···················· |
| R | 1257 | hCoV-19/Singapore/1199/2020\|EPI_ISL_648615\|2020-04 | ···················· | ···················· | ···················· | ···················· | ···················· | ···················· |
| R | 1258 | hCoV-19/Singapore/1200/2020\|EPI_ISL_648616\|2020-04 | ···················· | ···················· | ···················· | ···················· | ···················· | ···················· |
| R | 1259 | hCoV-19/Singapore/1201/2020\|EPI_ISL_648617\|2020-04 | ···················· | ···················· | ···················· | ···················· | ···················· | ···················· |
| R | 1260 | hCoV-19/Singapore/1202/2020\|EPI_ISL_648618\|2020-04 | ···················· | ···················· | ···················· | ···················· | ···················· | ···················· |
| R | 1261 | hCoV-19/Singapore/1203/2020\|EPI_ISL_648619\|2020-04 | ···················· | ···················· | ···················· | ···················· | ···················· | ···················· |
| R | 1262 | hCoV-19/Singapore/1204/2020\|EPI_ISL_648620\|2020-04 | ···················· | ···················· | ···················· | ···················· | ···················· | ···················· |
| R | 1263 | hCoV-19/Singapore/1205/2020\|EPI_ISL_648621\|2020-04 | ···················· | ···················· | ···················· | ···················· | ···················· | ···················· |
| R | 1264 | hCoV-19/Singapore/1206/2020\|EPI_ISL_648622\|2020-04 | ···················· | ···················· | ···················· | ···················· | ···················· | ···················· |
| R | 1265 | hCoV-19/Singapore/1207/2020\|EPI_ISL_648623\|2020-04 | ···················· | ···················· | ···················· | ···················· | ···················· | ···················· |
| R | 1266 | hCoV-19/Singapore/1208/2020\|EPI_ISL_648624\|2020-04 | ···················· | ···················· | ···················· | ···················· | ···················· | ···················· |
| R | 1267 | hCoV-19/Singapore/1209/2020\|EPI_ISL_648625\|2020-04 | ···················· | ···················· | ···················· | ···················· | ···················· | ···················· |
| R | 1268 | hCoV-19/Singapore/1210/2020\|EPI_ISL_648626\|2020-04 | ···················· | ···················· | ···················· | ···················· | ···················· | ···················· |
| R | 1269 | hCoV-19/Singapore/1211/2020\|EPI_ISL_648627\|2020-04 | ···················· | ···················· | ···················· | ···················· | ···················· | ···················· |
| R | 1270 | hCoV-19/Singapore/1212/2020\|EPI_ISL_648628\|2020-04 | ···················· | ···················· | ···················· | ···················· | ···················· | ···················· |
| R | 1271 | hCoV-19/Singapore/1213/2020\|EPI_ISL_648629\|2020-04 | ···················· | ···················· | ···················· | ···················· | ···················· | ···················· |
| R | 1272 | hCoV-19/Singapore/1214/2020\|EPI_ISL_648630\|2020-04 | ···················· | ···················· | ···················· | ···················· | ···················· | ···················· |
| R | 1273 | hCoV-19/Singapore/1215/2020\|EPI_ISL_648631\|2020-04 | ···················· | ···················· | ···················· | ···················· | ···················· | ···················· |
| R | 1274 | hCoV-19/Singapore/1216/2020\|EPI_ISL_648632\|2020-04 | ···················· | ···················· | ···················· | ···················· | ···················· | ···················· |
| R | 1275 | hCoV-19/Singapore/1217/2020\|EPI_ISL_648633\|2020-04 | ···················· | ···················· | ···················· | ···················· | ···················· | ···················· |
| R | 1276 | hCoV-19/Singapore/1218/2020\|EPI_ISL_648634\|2020-04 | ···················· | ···················· | ···················· | ···················· | ···················· | ···················· |
| R | 1277 | hCoV-19/Singapore/1219/2020\|EPI_ISL_648635\|2020-04 | ···················· | ···················· | ···················· | ···················· | ···················· | ···················· |
| R | 1278 | hCoV-19/Singapore/1220/2020\|EPI_ISL_648636\|2020-04 | ···················· | ···················· | ···················· | ···················· | ···················· | ···················· |
| R | 1279 | hCoV-19/Singapore/1221/2020\|EPI_ISL_648637\|2020-04 | ···················· | ···················· | ···················· | ···················· | ···················· | ···················· |

| | | | | | | | | |
|---|---|---|---|---|---|---|---|---|
| R | 1280 | hCoV-19/Singapore/1222/2020\|EPI_ISL_648638\|2020-04 | ···················· | ···················· | ····················· | ···················· | ···················· | ···················· |
| R | 1281 | hCoV-19/Australia/VIC11618/2020\|EPI_ISL_641000\|202 | ···················· | ···················· | ····················· | ···················· | ···················· | ···················· |
| R | 1282 | hCoV-19/Australia/VIC16064/2020\|EPI_ISL_641001\|202 | ···················· | ···················· | ····················· | ···················· | ···················· | ···················· |
| R | 1283 | hCoV-19/Australia/VIC11637/2020\|EPI_ISL_641003\|202 | ···················· | ···················· | ····················· | ···················· | ···················· | ···················· |
| R | 1284 | hCoV-19/Australia/VIC12166/2020\|EPI_ISL_641004\|202 | ···················· | ···················· | ····················· | ···················· | ···················· | ···················· |
| R | 1285 | hCoV-19/Australia/VIC12798/2020\|EPI_ISL_641005\|202 | ···················· | ···················· | ····················· | ···················· | ···················· | ···················· |
| R | 1286 | hCoV-19/Australia/VIC16068/2020\|EPI_ISL_641008\|202 | ···················· | ···················· | ····················· | ···················· | ···················· | ···················· |
| R | 1287 | hCoV-19/Australia/VIC16319/2020\|EPI_ISL_641009\|202 | ···················· | ···················· | ····················· | ···················· | ···················· | ···················· |
| R | 1288 | hCoV-19/Australia/VIC15929/2020\|EPI_ISL_641010\|202 | ···················· | ···················· | ····················· | ···················· | ···················· | ···················· |
| R | 1289 | hCoV-19/Australia/VIC12115/2020\|EPI_ISL_641011\|202 | ···················· | ···················· | ····················· | ···················· | ···················· | ···················· |
| R | 1292 | hCoV-19/Australia/VIC16359/2020\|EPI_ISL_641014\|202 | ···················· | ···················· | ····················· | ···················· | ···················· | ···················· |
| R | 1293 | hCoV-19/Australia/VIC16192/2020\|EPI_ISL_641017\|202 | ···················· | ···················· | ····················· | ···················· | ···················· | ···················· |
| R | 1294 | hCoV-19/Australia/VIC13269/2020\|EPI_ISL_641018\|202 | ···················· | ···················· | ····················· | ···················· | ···················· | ···················· |
| R | 1295 | hCoV-19/Australia/VIC12206/2020\|EPI_ISL_641020\|202 | ···················· | ···················· | ····················· | ···················· | ···················· | ···················· |
| R | 1297 | hCoV-19/Australia/VIC13124/2020\|EPI_ISL_641022\|202 | ···················· | ···················· | ····················· | ···················· | ···················· | ···················· |
| R | 1298 | hCoV-19/Australia/VIC16046/2020\|EPI_ISL_641024\|202 | ···················· | ···················· | ····················· | ···················· | ···················· | ···················· |
| R | 1299 | hCoV-19/Australia/VIC12106/2020\|EPI_ISL_641025\|202 | ···················· | ···················· | ····················· | ···················· | ···················· | ···················· |
| R | 1301 | hCoV-19/Australia/VIC12360/2020\|EPI_ISL_641027\|202 | ···················· | ···················· | ····················· | ···················· | ···················· | ···················· |
| R | 1303 | hCoV-19/Australia/VIC11645/2020\|EPI_ISL_641029\|202 | ···················· | ···················· | ····················· | ···················· | ···················· | ···················· |
| R | 1304 | hCoV-19/Australia/VIC13150/2020\|EPI_ISL_641030\|202 | ···················· | ···················· | ····················· | ···················· | ···················· | ···················· |
| R | 1305 | hCoV-19/Australia/VIC12507/2020\|EPI_ISL_641032\|202 | ···················· | ···················· | ····················· | ···················· | ···················· | ···················· |
| R | 1306 | hCoV-19/Australia/VIC12071/2020\|EPI_ISL_641034\|202 | ···················· | ···················· | ····················· | ···················· | ···················· | ···················· |
| R | 1307 | hCoV-19/Australia/VIC12118/2020\|EPI_ISL_641035\|202 | ···················· | ···················· | ····················· | ···················· | ···················· | ···················· |
| R | 1308 | hCoV-19/Australia/VIC13022/2020\|EPI_ISL_641036\|202 | ···················· | ···················· | ····················· | ···················· | ···················· | ···················· |
| R | 1309 | hCoV-19/Australia/VIC15965/2020\|EPI_ISL_641038\|202 | ···················· | ···················· | ····················· | ···················· | ···················· | ···················· |
| R | 1310 | hCoV-19/Australia/VIC12437/2020\|EPI_ISL_641040\|202 | ···················· | ···················· | ····················· | ···················· | ···················· | ···················· |
| R | 1311 | hCoV-19/Australia/VIC15888/2020\|EPI_ISL_641041\|202 | ···················· | ···················· | ····················· | ···················· | ···················· | ···················· |
| R | 1312 | hCoV-19/Australia/VIC12435/2020\|EPI_ISL_641042\|202 | ···················· | ···················· | ····················· | ···················· | ···················· | ···················· |
| R | 1313 | hCoV-19/Australia/VIC12485/2020\|EPI_ISL_641043\|202 | ···················· | ···················· | ····················· | ···················· | ···················· | ···················· |
| R | 1314 | hCoV-19/Australia/VIC11981/2020\|EPI_ISL_641045\|202 | ···················· | ···················· | ····················· | ···················· | ···················· | ···················· |
| R | 1315 | hCoV-19/Australia/VIC12027/2020\|EPI_ISL_641046\|202 | ···················· | ···················· | ····················· | ···················· | ···················· | ···················· |
| R | 1316 | hCoV-19/Australia/VIC16257/2020\|EPI_ISL_641048\|202 | ···················· | ···················· | ····················· | ···················· | ···················· | ···················· |
| R | 1317 | hCoV-19/Australia/VIC12744/2020\|EPI_ISL_641071\|202 | ···················· | ···················· | ····················· | ···················· | ···················· | ···················· |
| R | 1318 | hCoV-19/Australia/VIC12284/2020\|EPI_ISL_641073\|202 | ···················· | ···················· | ····················· | ···················· | ···················· | ···················· |
| R | 1319 | hCoV-19/Australia/VIC16061/2020\|EPI_ISL_641074\|202 | ···················· | ···················· | ····················· | ···················· | ···················· | ···················· |
| R | 1320 | hCoV-19/Australia/VIC16183/2020\|EPI_ISL_641077\|202 | ···················· | ···················· | ····················· | ···················· | ···················· | ···················· |
| R | 1321 | hCoV-19/Australia/VIC16444/2020\|EPI_ISL_641078\|202 | ···················· | ···················· | ····················· | ···················· | ···················· | ···················· |
| R | 1322 | hCoV-19/Australia/VIC12579/2020\|EPI_ISL_641082\|202 | ···················· | ···················· | ····················· | ···················· | ···················· | ···················· |
| R | 1323 | hCoV-19/Australia/VIC16171/2020\|EPI_ISL_641083\|202 | ···················· | ···················· | ····················· | ···················· | ···················· | ···················· |
| R | 1324 | hCoV-19/Australia/VIC12760/2020\|EPI_ISL_641084\|202 | ···················· | ···················· | ····················· | ···················· | ···················· | ···················· |
| R | 1325 | hCoV-19/Australia/VIC13089/2020\|EPI_ISL_641093\|202 | ···················· | ···················· | ····················· | ···················· | ···················· | ···················· |
| R | 1326 | hCoV-19/Australia/VIC15935/2020\|EPI_ISL_641094\|202 | ···················· | ···················· | ····················· | ···················· | ···················· | ···················· |
| R | 1327 | hCoV-19/Australia/VIC13056/2020\|EPI_ISL_641095\|202 | ···················· | ···················· | ····················· | ···················· | ···················· | ···················· |
| R | 1328 | hCoV-19/Australia/VIC12022/2020\|EPI_ISL_641096\|202 | ···················· | ···················· | ····················· | ···················· | ···················· | ···················· |
| R | 1329 | hCoV-19/Australia/VIC12141/2020\|EPI_ISL_641099\|202 | ···················· | ···················· | ····················· | ···················· | ···················· | ···················· |
| R | 1330 | hCoV-19/England/QEUH-B1155D/2020\|EPI_ISL_642301\|20 | ···················· | ···················· | ····················· | ···················· | ···················· | ···················· |
| R | 1331 | hCoV-19/England/QEUH-B12176/2020\|EPI_ISL_642302\|20 | ···················· | ···················· | ····················· | ···················· | ···················· | ···················· |
| R | 1332 | hCoV-19/England/QEUH-B121FE/2020\|EPI_ISL_642303\|20 | ···················· | ···················· | ····················· | ···················· | ···················· | ···················· |
| R | 1333 | hCoV-19/England/QEUH-B10CB3/2020\|EPI_ISL_642304\|20 | ···················· | ·A·················· | ····················· | ···················· | ···················· | ···················· |
| R | 1334 | hCoV-19/England/QEUH-B116F0/2020\|EPI_ISL_642305\|20 | ···················· | ···················· | ····················· | ···················· | ···················· | ···················· |
| R | 1335 | hCoV-19/England/QEUH-B11A8B/2020\|EPI_ISL_642306\|20 | ···················· | ···················· | ····················· | ···················· | ···················· | ···················· |
| R | 1336 | hCoV-19/England/QEUH-B1199D/2020\|EPI_ISL_642307\|20 | ···················· | ···················· | ····················· | ···················· | ···················· | ···················· |
| R | 1337 | hCoV-19/England/QEUH-B11CA3/2020\|EPI_ISL_642308\|20 | ···················· | ···················· | ····················· | ···················· | ···················· | ···················· |
| R | 1338 | hCoV-19/England/QEUH-B11A7C/2020\|EPI_ISL_642309\|20 | ···················· | ···················· | ····················· | ···················· | ···················· | ···················· |
| R | 1339 | hCoV-19/England/QEUH-B10E35/2020\|EPI_ISL_642310\|20 | ···················· | ···················· | ····················· | ···················· | ···················· | ···················· |
| R | 1340 | hCoV-19/England/QEUH-B128F9/2020\|EPI_ISL_642311\|20 | ···················· | ···················· | ····················· | ···················· | ···················· | ···················· |
| R | 1341 | hCoV-19/England/QEUH-B1048E/2020\|EPI_ISL_642312\|20 | ···················· | ···················· | ····················· | ···················· | ···················· | ···················· |
| R | 1342 | hCoV-19/England/QEUH-B13649/2020\|EPI_ISL_642313\|20 | ···················· | ···················· | ····················· | ···················· | ···················· | ···················· |
| R | 1343 | hCoV-19/England/QEUH-B116C3/2020\|EPI_ISL_642314\|20 | ···················· | ···················· | ····················· | ···················· | ···················· | ···················· |
| R | 1344 | hCoV-19/England/QEUH-B11872/2020\|EPI_ISL_642315\|20 | ···················· | ···················· | ····················· | ···················· | ···················· | ···················· |
| R | 1345 | hCoV-19/England/QEUH-B1310C/2020\|EPI_ISL_642316\|20 | ···················· | ···················· | ····················· | ···················· | ···················· | ···················· |
| R | 1346 | hCoV-19/England/QEUH-B10E44/2020\|EPI_ISL_642318\|20 | ···················· | ···················· | ····················· | ···················· | ···················· | ···················· |
| R | 1347 | hCoV-19/England/QEUH-B121B2/2020\|EPI_ISL_642324\|20 | ···················· | ···················· | ····················· | ···················· | ···················· | ···················· |
| R | 1348 | hCoV-19/England/QEUH-B117FD/2020\|EPI_ISL_642325\|20 | ···················· | ···················· | ····················· | ···················· | ···················· | ···················· |
| R | 1349 | hCoV-19/England/QEUH-B104E8/2020\|EPI_ISL_642326\|20 | ···················· | ···················· | ····················· | ···················· | ···················· | ···················· |
| R | 1350 | hCoV-19/England/QEUH-B11B1F/2020\|EPI_ISL_642327\|20 | ···················· | ·A·················· | ····················· | ···················· | ···················· | ···················· |
| R | 1351 | hCoV-19/England/QEUH-B13218/2020\|EPI_ISL_642328\|20 | ···················· | ···················· | ····················· | ···················· | ···················· | ···················· |
| R | 1352 | hCoV-19/England/QEUH-B11C2B/2020\|EPI_ISL_642329\|20 | ···················· | ···················· | ····················· | ···················· | ···················· | ···················· |
| R | 1353 | hCoV-19/England/QEUH-B117B1/2020\|EPI_ISL_642330\|20 | ···················· | ···················· | ····················· | ···················· | ···················· | ···················· |
| R | 1354 | hCoV-19/England/QEUH-B11D28/2020\|EPI_ISL_642331\|20 | ···················· | ···················· | ····················· | ···················· | ···················· | ···················· |
| R | 1355 | hCoV-19/England/QEUH-B11BA6/2020\|EPI_ISL_642332\|20 | ···················· | ···················· | ····················· | ···················· | ·····T·············· | ···················· |
| R | 1356 | hCoV-19/England/QEUH-B11933/2020\|EPI_ISL_642333\|20 | ···················· | ···················· | ····················· | ···················· | ···················· | ···················· |
| R | 1357 | hCoV-19/England/QEUH-B127B0/2020\|EPI_ISL_642334\|20 | ···················· | ···················· | ····················· | ···················· | ···················· | ···················· |
| R | 1358 | hCoV-19/England/QEUH-B10EDB/2020\|EPI_ISL_642335\|20 | ···················· | ···················· | ····················· | ···················· | ···················· | ···················· |
| R | 1359 | hCoV-19/England/QEUH-B13694/2020\|EPI_ISL_642336\|20 | ···················· | ···················· | ····················· | ···················· | ···················· | ···················· |
| R | 1360 | hCoV-19/England/QEUH-B12880/2020\|EPI_ISL_642337\|20 | ···················· | ···················· | ····················· | ···················· | ···················· | ···················· |
| R | 1361 | hCoV-19/England/QEUH-B117DF/2020\|EPI_ISL_642338\|20 | ···················· | ···················· | ····················· | ···················· | ···················· | ···················· |
| R | 1362 | hCoV-19/England/QEUH-B10CFF/2020\|EPI_ISL_642339\|20 | ···················· | ···················· | ····················· | ···················· | ···················· | ···················· |
| R | 1363 | hCoV-19/England/QEUH-B10F41/2020\|EPI_ISL_642340\|20 | ···················· | ···················· | ····················· | ···················· | ···················· | ···················· |
| R | 1364 | hCoV-19/England/QEUH-B12853/2020\|EPI_ISL_642341\|20 | ···················· | ···················· | ····················· | ···················· | ···················· | ···················· |

R 1365 hCoV-19/England/QEUH-B10E26/2020|EPI_ISL_642342|20 ···················· ···················· ···················· ···················· ···················· ····················
R 1366 hCoV-19/England/QEUH-B1057C/2020|EPI_ISL_642343|20 ···················· ···················· ···················· ···················· ···················· ····················
R 1367 hCoV-19/England/QEUH-B1289F/2020|EPI_ISL_642345|20 ···················· ···················· ···················· ···················· ···················· ····················
R 1368 hCoV-19/England/QEUH-B115F3/2020|EPI_ISL_642346|20 ···················· ···················· ···················· ···················· ···················· ····················
R 1369 hCoV-19/England/QEUH-B1220A/2020|EPI_ISL_642347|20 ···················· ···················· ···················· ···················· ···················· ····················
R 1370 hCoV-19/England/QEUH-B10D74/2020|EPI_ISL_642348|20 ···················· ···················· ···················· ···················· ···················· ····················
R 1371 hCoV-19/England/QEUH-B104BB/2020|EPI_ISL_642349|20 ···················· ···················· ···················· ···················· ···················· ····················
R 1372 hCoV-19/England/QEUH-B11450/2020|EPI_ISL_642350|20 ···················· ·A·················· ···················· ···················· ···················· ····················
R 1373 hCoV-19/England/QEUH-B11CEF/2020|EPI_ISL_642351|20 ···················· ···················· ···················· ···················· ···················· ····················
R 1374 hCoV-19/England/QEUH-B11775/2020|EPI_ISL_642352|20 ···················· ···················· ···················· ···················· ···················· ····················
R 1375 hCoV-19/England/QEUH-B12149/2020|EPI_ISL_642353|20 ···················· ···················· ···················· ···················· ···················· ····················
R 1376 hCoV-19/England/QEUH-B13087/2020|EPI_ISL_642354|20 ···················· ···················· ···················· ···················· ···················· ····················
R 1377 hCoV-19/England/QEUH-B11678/2020|EPI_ISL_642355|20 ···················· ···················· ···················· ···················· ···················· ····················
R 1378 hCoV-19/England/QEUH-B1156C/2020|EPI_ISL_642356|20 ···················· ···················· ···················· ···················· ···················· ····················
R 1379 hCoV-19/England/QEUH-B122A0/2020|EPI_ISL_642357|20 ···················· ···················· ···················· ···················· ···················· ····················
R 1380 hCoV-19/England/QEUH-B114D8/2020|EPI_ISL_642358|20 ···················· ·A·················· ···················· ···················· ···················· ····················
R 1381 hCoV-19/England/QEUH-B12862/2020|EPI_ISL_642359|20 ···················· ···················· ···················· ···················· ···················· ····················
R 1382 hCoV-19/England/QEUH-B10E9F/2020|EPI_ISL_642360|20 ···················· ···················· ···················· ···················· ···················· ····················
R 1383 hCoV-19/England/QEUH-B131EE/2020|EPI_ISL_642362|20 ···················· ···················· ···················· ···················· ···················· ····················
R 1384 hCoV-19/England/QEUH-B1158A/2020|EPI_ISL_642364|20 ···················· ···················· ···················· ···················· ···················· ····················
R 1385 hCoV-19/England/QEUH-B11836/2020|EPI_ISL_642365|20 ···················· ···················· ···················· ···················· ···················· ····················
R 1386 hCoV-19/England/QEUH-B130F0/2020|EPI_ISL_642366|20 ···················· ···················· ···················· ···················· ···················· ····················
R 1387 hCoV-19/England/QEUH-B10E62/2020|EPI_ISL_642367|20 ···················· ···················· ···················· ···················· ···················· ····················
R 1388 hCoV-19/England/QEUH-B104AC/2020|EPI_ISL_642368|20 ···················· ···················· ···················· ···················· ···················· ····················
R 1389 hCoV-19/England/QEUH-B11C3A/2020|EPI_ISL_642369|20 ···················· ·A·················· ···················· ···················· ···················· ···A················
R 1390 hCoV-19/England/QEUH-B118BE/2020|EPI_ISL_642370|20 ···················· ···················· ···················· ···················· ···················· ····················
R 1391 hCoV-19/England/QEUH-B12774/2020|EPI_ISL_642371|20 ···················· ···················· ···················· ···················· ···················· ····················
R 1392 hCoV-19/England/QEUH-B10D29/2020|EPI_ISL_642373|20 ···················· ···················· ···················· ···················· ···················· ····················
R 1393 hCoV-19/England/QEUH-B104D9/2020|EPI_ISL_642374|20 ···················· ···················· ···················· ···················· ···················· ····················
R 1394 hCoV-19/England/QEUH-B118CD/2020|EPI_ISL_642375|20 ···················· ···················· ···················· ···················· ···················· ····················
R 1395 hCoV-19/England/QEUH-B11DEC/2020|EPI_ISL_642376|20 ···················· ···················· ···················· ···················· ···················· ····················
R 1396 hCoV-19/England/QEUH-B115A8/2020|EPI_ISL_642377|20 ···················· ···················· ···················· ···················· ···················· ····················
R 1397 hCoV-19/England/QEUH-B1311B/2020|EPI_ISL_642379|20 ···················· ···················· ···················· ···················· ···················· ····················
R 1398 hCoV-19/England/CAMC-B080D0/2020|EPI_ISL_643623|20 ···················· ···················· ···················· ···················· ···················· ····················
R 1399 hCoV-19/England/CAMC-B0AD22/2020|EPI_ISL_643625|20 ···················· ···················· ···················· ···················· ···················· ····················
R 1400 hCoV-19/England/CAMC-B0CC05/2020|EPI_ISL_643627|20 ···················· ···················· ···················· ···················· ···················· ····················
R 1401 hCoV-19/England/CAMC-B0C6BC/2020|EPI_ISL_643628|20 ···················· ···················· ···················· ···················· ······T············· ····················
R 1402 hCoV-19/England/CAMC-B0BAC0/2020|EPI_ISL_643629|20 ···················· ···················· ···················· ···················· ···················· ····················
R 1403 hCoV-19/England/CAMC-B0AA94/2020|EPI_ISL_643630|20 ···················· ···················· ···················· ···················· ···················· ····················
R 1404 hCoV-19/England/CAMC-B07E9B/2020|EPI_ISL_643631|20 ···················· ···················· ···················· ···················· ···················· ····················
R 1405 hCoV-19/England/CAMC-B0ADF5/2020|EPI_ISL_643632|20 ···················· ···················· ···················· ···················· ···················· ····················
R 1406 hCoV-19/England/CAMC-B0B6F9/2020|EPI_ISL_643633|20 ···················· ···················· ···················· ···················· ···················· ····················
R 1407 hCoV-19/England/CAMC-B0AEA7/2020|EPI_ISL_643635|20 ···················· ···················· ···················· ···················· ···················· ····················
R 1408 hCoV-19/England/CAMC-B0AF68/2020|EPI_ISL_643636|20 ···················· ···················· ···················· ···················· ···················· ····················
R 1409 hCoV-19/England/CAMC-B082BC/2020|EPI_ISL_643637|20 ···················· ···················· ···················· ···················· ···················· ····················
R 1410 hCoV-19/England/CAMC-B083E6/2020|EPI_ISL_643638|20 ···················· ···················· ···················· ···················· ···················· ····················
R 1411 hCoV-19/England/CAMC-B08CAF/2020|EPI_ISL_643639|20 ···················· ···················· ···················· ···················· ···················· ····················
R 1412 hCoV-19/England/CAMC-B08814/2020|EPI_ISL_643640|20 ···················· ···················· ···················· ···················· ···················· ····················
R 1413 hCoV-19/England/CAMC-B07D61/2020|EPI_ISL_643642|20 ···················· ···················· ···················· ···················· ···················· ····················
R 1414 hCoV-19/England/CAMC-B07FA7/2020|EPI_ISL_643643|20 ···················· ···················· ···················· ···················· ···················· ····················
R 1415 hCoV-19/England/CAMC-B08313/2020|EPI_ISL_643644|20 ···················· ···················· ···················· ···················· ···················· ····················
R 1416 hCoV-19/England/CAMC-B07A00/2020|EPI_ISL_643645|20 ···················· ···················· ···················· ···················· ···················· ····················
R 1417 hCoV-19/England/CAMC-B0888D/2020|EPI_ISL_643646|20 ···················· ·A·················· ···················· ···················· ···················· ···A················
R 1418 hCoV-19/England/CAMC-B07842/2020|EPI_ISL_643647|20 ···················· ···················· ···················· ···················· ···················· ····················
R 1419 hCoV-19/England/CAMC-B087F9/2020|EPI_ISL_643648|20 ···················· ···················· ···················· ···················· ···················· ····················
R 1420 hCoV-19/England/CAMC-B078BB/2020|EPI_ISL_643649|20 ···················· ···················· ···················· ···················· ···················· ····················
R 1421 hCoV-19/England/CAMC-B074C6/2020|EPI_ISL_643650|20 ···················· ···················· ···················· ···················· ···················· ····················
R 1422 hCoV-19/England/CAMC-B08146/2020|EPI_ISL_643651|20 ···················· ···················· ···················· ···················· ···················· ····················
R 1423 hCoV-19/England/CAMC-B09251/2020|EPI_ISL_643652|20 ···················· ···················· ···················· ···················· ···················· ····················
R 1424 hCoV-19/England/CAMC-B0711A/2020|EPI_ISL_643653|20 ···················· ·A·················· ···················· ···················· ···················· ···A················
R 1425 hCoV-19/England/CAMC-B07F7A/2020|EPI_ISL_643654|20 ···················· ···················· ···················· ···················· ···················· ····················
R 1426 hCoV-19/England/CAMC-B073BA/2020|EPI_ISL_643657|20 ···················· ···················· ···················· ···················· ···················· ····················
R 1427 hCoV-19/England/CAMC-B0710B/2020|EPI_ISL_643658|20 ···················· ···················· ···················· ···················· ···················· ····················
R 1428 hCoV-19/England/CAMC-B07903/2020|EPI_ISL_643659|20 ···················· ···················· ···················· ···················· ···················· ····················
R 1429 hCoV-19/England/CAMC-B0753C/2020|EPI_ISL_643660|20 ···················· ···················· ···················· ···················· ···················· ····················
R 1430 hCoV-19/England/CAMC-B07411/2020|EPI_ISL_643661|20 ···················· ···················· ···················· ···················· ······T············· ····················
R 1431 hCoV-19/England/CAMC-B07B94/2020|EPI_ISL_643662|20 ···················· ···················· ···················· ···················· ······T············· ····················
R 1432 hCoV-19/England/CAMC-B0893F/2020|EPI_ISL_643663|20 ···················· ·R·················· ···················· ···················· ···················· ···A················
R 1433 hCoV-19/England/CAMC-B08780/2020|EPI_ISL_643664|20 ···················· ···················· ···················· ···················· ···················· ····················
R 1434 hCoV-19/England/CAMC-B07208/2020|EPI_ISL_643665|20 ···················· ·A·················· ···················· ···················· ···················· ···A················
R 1435 hCoV-19/England/CAMC-B0795E/2020|EPI_ISL_643667|20 ···················· ···················· ···················· ···················· ···················· ····················
R 1436 hCoV-19/England/CAMC-B08771/2020|EPI_ISL_643668|20 ···················· ···················· ···················· ···················· ···················· ····················
R 1437 hCoV-19/England/CAMC-B07727/2020|EPI_ISL_643669|20 ···················· ·A·················· ···················· ···················· ···················· ···A················
R 1438 hCoV-19/England/CAMC-B0797C/2020|EPI_ISL_643670|20 ···················· ···················· ···················· ···················· ···················· ····················
R 1439 hCoV-19/England/CAMC-B08841/2020|EPI_ISL_643671|20 ···················· ···················· ···················· ···················· ······T············· ····················
R 1440 hCoV-19/England/CAMC-B0836E/2020|EPI_ISL_643672|20 ···················· ···················· ···················· ···················· ···················· ····················
R 1441 hCoV-19/England/CAMC-B07174/2020|EPI_ISL_643673|20 ···················· ···················· ···················· ···················· ···················· ····················
R 1442 hCoV-19/England/CAMC-B08CCD/2020|EPI_ISL_643674|20 ···················· ·A·······A·········· ···················· ···················· ···················· ····················
R 1443 hCoV-19/England/CAMC-B07BD0/2020|EPI_ISL_643675|20 ···················· ···················· ···················· ···················· ······T············· ····················
R 1444 hCoV-19/England/CAMC-B07C82/2020|EPI_ISL_643677|20 ···················· ···················· ···················· ···················· ···················· ····················

| | | | | | | | |
|---|---|---|---|---|---|---|---|
| R | 1445 hCoV-19/England/CAMC-B088AB/2020\|EPI_ISL_643678\|20 | ···················· | ·A·················· | ····················· | ···················· | ···················· | ··A················· |
| R | 1446 hCoV-19/England/CAMC-B07C46/2020\|EPI_ISL_643679\|20 | ···················· | ···················· | ····················· | ···················· | ···················· | ···················· |
| R | 1447 hCoV-19/England/CAMC-B0762A/2020\|EPI_ISL_643680\|20 | ···················· | ···················· | ····················· | ···················· | ···················· | ···················· |
| R | 1448 hCoV-19/England/CAMC-B08656/2020\|EPI_ISL_643681\|20 | ···················· | ···················· | ····················· | ···················· | ···················· | ···················· |
| R | 1449 hCoV-19/England/CAMC-B07578/2020\|EPI_ISL_643683\|20 | ···················· | ···················· | ····················· | ···················· | ·····T·············· | ···················· |
| R | 1450 hCoV-19/England/CAMC-B073F6/2020\|EPI_ISL_643685\|20 | ···················· | ···················· | ····················· | ···················· | ···················· | ···················· |
| R | 1451 hCoV-19/England/CAMC-B0835F/2020\|EPI_ISL_643686\|20 | ···················· | ···················· | ····················· | ···················· | ···················· | ···················· |
| R | 1452 hCoV-19/England/CAMC-B08902/2020\|EPI_ISL_643687\|20 | ···················· | ···················· | ····················· | ···················· | ···················· | ···················· |
| R | 1453 hCoV-19/England/CAMC-B079F4/2020\|EPI_ISL_643688\|20 | ···················· | ···················· | ····················· | ···················· | ···················· | ···················· |
| R | 1454 hCoV-19/England/CAMC-B07138/2020\|EPI_ISL_643689\|20 | ···················· | ···················· | ····················· | ···················· | ···················· | ···················· |
| R | 1455 hCoV-19/England/CAMC-B08401/2020\|EPI_ISL_643690\|20 | ···················· | ·A·················· | ····················· | ···················· | ···················· | ··A················· |
| R | 1456 hCoV-19/England/CAMC-B08AD2/2020\|EPI_ISL_643691\|20 | ···················· | ···················· | ····················· | ···················· | ···················· | ···················· |
| R | 1457 hCoV-19/England/CAMC-B07BA3/2020\|EPI_ISL_643692\|20 | ···················· | ···················· | ····················· | ···················· | ·····T·············· | ···················· |
| R | 1458 hCoV-19/England/CAMC-B07F5C/2020\|EPI_ISL_643693\|20 | ···················· | ···················· | ····················· | ···················· | ···················· | ···················· |
| R | 1459 hCoV-19/England/CAMC-B072AE/2020\|EPI_ISL_643694\|20 | ···················· | ···················· | ····················· | ···················· | ···················· | ···················· |
| R | 1460 hCoV-19/England/CAMC-B07EC8/2020\|EPI_ISL_643695\|20 | ···················· | ···················· | ····················· | ···················· | ···················· | ···················· |
| R | 1461 hCoV-19/England/CAMC-B081DD/2020\|EPI_ISL_643696\|20 | ···················· | ···················· | ····················· | ···················· | ···················· | ···················· |
| R | 1462 hCoV-19/England/CAMC-B081A0/2020\|EPI_ISL_643699\|20 | ···················· | ···················· | ····················· | ···················· | ···················· | ···················· |
| R | 1463 hCoV-19/Japan/Donner49/2020\|EPI_ISL_644954\|2020-06 | ···················· | ···················· | ····················· | ···················· | ···················· | ···················· |
| R | 1464 hCoV-19/Japan/Donner50/2020\|EPI_ISL_644955\|2020-07 | ···················· | ···················· | ····················· | ···················· | ···················· | ···················· |
| R | 1465 hCoV-19/Japan/Donner51/2020\|EPI_ISL_644956\|2020-07 | ···················· | ···················· | ····················· | ···················· | ···················· | ···················· |
| R | 1466 hCoV-19/Japan/Donner52/2020\|EPI_ISL_644957\|2020-07 | ···················· | ···················· | ····················· | ···················· | ···················· | ···················· |
| R | 1467 hCoV-19/Japan/Donner53/2020\|EPI_ISL_644958\|2020-07 | ···················· | ···················· | ····················· | ···················· | ···················· | ···················· |
| R | 1468 hCoV-19/Japan/Donner54/2020\|EPI_ISL_644959\|2020-07 | ···················· | ···················· | ····················· | ···················· | ···················· | ···················· |
| R | 1469 hCoV-19/Japan/Donner55/2020\|EPI_ISL_644960\|2020-07 | ···················· | ···················· | ····················· | ···················· | ···················· | ···················· |
| R | 1470 hCoV-19/Japan/Donner57/2020\|EPI_ISL_644961\|2020-07 | ···················· | ···················· | ····················· | ···················· | ···················· | ···················· |
| R | 1471 hCoV-19/Japan/Donner58/2020\|EPI_ISL_644962\|2020-07 | ···················· | ···················· | ····················· | ···················· | ···················· | ···················· |
| R | 1472 hCoV-19/Japan/Donner59/2020\|EPI_ISL_644963\|2020-07 | ···················· | ···················· | ····················· | ···················· | ···················· | ···················· |
| R | 1473 hCoV-19/Japan/Donner60/2020\|EPI_ISL_644964\|2020-07 | ···················· | ···················· | ····················· | ···················· | ···················· | ···················· |
| R | 1474 hCoV-19/Japan/Donner61/2020\|EPI_ISL_644965\|2020-07 | ···················· | ···················· | ····················· | ···················· | ···················· | ···················· |
| R | 1475 hCoV-19/Japan/Donner62/2020\|EPI_ISL_644966\|2020-07 | ···················· | ···················· | ····················· | ···················· | ···················· | ···················· |
| R | 1476 hCoV-19/Japan/Donner63/2020\|EPI_ISL_644967\|2020-07 | ···················· | ···················· | ····················· | ···················· | ···················· | ···················· |
| R | 1477 hCoV-19/Japan/Donner64/2020\|EPI_ISL_644968\|2020-07 | ···················· | ···················· | ····················· | ···················· | ···················· | ···················· |
| R | 1478 hCoV-19/Japan/Donner65/2020\|EPI_ISL_644969\|2020-07 | ···················· | ···················· | ····················· | ···················· | ···················· | ···················· |
| R | 1479 hCoV-19/Japan/Donner66/2020\|EPI_ISL_644970\|2020-07 | ···················· | ···················· | ····················· | ···················· | ···················· | ···················· |
| R | 1480 hCoV-19/Japan/Donner67/2020\|EPI_ISL_644971\|2020-07 | ···················· | ···················· | ····················· | ···················· | ···················· | ···················· |
| R | 1481 hCoV-19/Japan/Donner68/2020\|EPI_ISL_644972\|2020-07 | ···················· | ···················· | ····················· | ···················· | ···················· | ···················· |
| R | 1482 hCoV-19/Japan/Donner69/2020\|EPI_ISL_644973\|2020-07 | ···················· | ···················· | ····················· | ···················· | ···················· | ···················· |
| R | 1483 hCoV-19/Japan/Donner70/2020\|EPI_ISL_644974\|2020-07 | ···················· | ···················· | ····················· | ···················· | ···················· | ···················· |
| R | 1484 hCoV-19/Japan/Donner71/2020\|EPI_ISL_644975\|2020-07 | ···················· | ···················· | ····················· | ···················· | ···················· | ···················· |
| R | 1485 hCoV-19/Japan/Donner72/2020\|EPI_ISL_644976\|2020-07 | ···················· | ···················· | ····················· | ···················· | ···················· | ···················· |
| R | 1486 hCoV-19/Japan/Donner73/2020\|EPI_ISL_644977\|2020-07 | ···················· | ···················· | ····················· | ···················· | ···················· | ···················· |
| R | 1487 hCoV-19/Japan/Donner74/2020\|EPI_ISL_644978\|2020-07 | ···················· | ···················· | ····················· | ···················· | ···················· | ···················· |
| R | 1488 hCoV-19/Japan/Donner75/2020\|EPI_ISL_644979\|2020-07 | ···················· | ···················· | ····················· | ···················· | ···················· | ···················· |
| R | 1489 hCoV-19/Japan/Donner76/2020\|EPI_ISL_644980\|2020-07 | ···················· | ···················· | ····················· | ···················· | ···················· | ···················· |
| R | 1490 hCoV-19/Japan/Donner77/2020\|EPI_ISL_644981\|2020-08 | ···················· | ···················· | ····················· | ···················· | ···················· | ···················· |
| R | 1491 hCoV-19/Japan/Donner78/2020\|EPI_ISL_644982\|2020-08 | ···················· | ···················· | ····················· | ···················· | ···················· | ···················· |
| R | 1492 hCoV-19/Japan/Donner79/2020\|EPI_ISL_644983\|2020-08 | ···················· | ···················· | ····················· | ···················· | ···················· | ···················· |
| R | 1493 hCoV-19/Japan/Donner80/2020\|EPI_ISL_644984\|2020-08 | ···················· | ···················· | ····················· | ···················· | ···················· | ···················· |
| R | 1494 hCoV-19/Japan/Donner81/2020\|EPI_ISL_644985\|2020-08 | ···················· | ···················· | ····················· | ···················· | ···················· | ···················· |
| R | 1495 hCoV-19/Japan/Donner82/2020\|EPI_ISL_644986\|2020-08 | ···················· | ···················· | ····················· | ···················· | ···················· | ···················· |
| R | 1496 hCoV-19/Japan/Donner83/2020\|EPI_ISL_644987\|2020-08 | ···················· | ···················· | ····················· | ···················· | ···················· | ···················· |
| R | 1497 hCoV-19/Japan/Donner84/2020\|EPI_ISL_644988\|2020-08 | ···················· | ···················· | ····················· | ···················· | ···················· | ···················· |
| R | 1498 hCoV-19/Japan/Donner85/2020\|EPI_ISL_644989\|2020-08 | ···················· | ···················· | ····················· | ···················· | ···················· | ···················· |
| R | 1499 hCoV-19/Japan/Donner86/2020\|EPI_ISL_644990\|2020-08 | ···················· | ···················· | ····················· | ···················· | ···················· | ···················· |
| R | 1500 hCoV-19/Japan/Donner87/2020\|EPI_ISL_644991\|2020-08 | ···················· | ···················· | ····················· | ···················· | ···················· | ···················· |
| R | 1501 hCoV-19/Japan/Donner88/2020\|EPI_ISL_644992\|2020-08 | ···················· | ···················· | ····················· | ···················· | ···················· | ···················· |
| R | 1502 hCoV-19/Japan/Donner89/2020\|EPI_ISL_644993\|2020-08 | ···················· | ···················· | ····················· | ···················· | ···················· | ···················· |
| R | 1503 hCoV-19/Japan/Donner90/2020\|EPI_ISL_644994\|2020-08 | ···················· | ···················· | ····················· | ···················· | ···················· | ···················· |
| R | 1504 hCoV-19/Japan/Donner91/2020\|EPI_ISL_644995\|2020-08 | ········T··········· | ···················· | ····················· | ···················· | ···················· | ···················· |
| R | 1505 hCoV-19/Japan/Donner92/2020\|EPI_ISL_644996\|2020-08 | ···················· | ···················· | ····················· | ···················· | ···················· | ···················· |
| R | 1506 hCoV-19/Japan/Donner93/2020\|EPI_ISL_644997\|2020-08 | ···················· | ···················· | ····················· | ···················· | ···················· | ···················· |
| R | 1507 hCoV-19/Japan/Donner94/2020\|EPI_ISL_644998\|2020-08 | ···················· | ···················· | ····················· | ···················· | ···················· | ···················· |
| R | 1508 hCoV-19/Japan/Donner95/2020\|EPI_ISL_644999\|2020-08 | ···················· | ···················· | ····················· | ···················· | ···················· | ···················· |
| R | 1509 hCoV-19/England/CAMC-B224C5/2020\|EPI_ISL_647270\|20 | ···················· | ···················· | ····················· | ···················· | ···················· | ···················· |
| R | 1510 hCoV-19/England/CAMC-B21763/2020\|EPI_ISL_647271\|20 | ···················· | ···················· | ····················· | ···················· | ···················· | ···················· |
| R | 1511 hCoV-19/England/CAMC-B221DD/2020\|EPI_ISL_647272\|20 | ···················· | ···················· | ····················· | ···················· | ···················· | ···················· |
| R | 1512 hCoV-19/England/CAMC-B21912/2020\|EPI_ISL_647273\|20 | ···················· | ···················· | ····················· | ···················· | ···················· | ···················· |
| R | 1513 hCoV-19/England/CAMC-B21F98/2020\|EPI_ISL_647274\|20 | ···················· | ···················· | ····················· | ···················· | ···················· | ···················· |
| R | 1514 hCoV-19/England/CAMC-B22DF7/2020\|EPI_ISL_647275\|20 | ···················· | ···················· | ····················· | ···················· | ···················· | ···················· |
| R | 1515 hCoV-19/England/CAMC-B22D42/2020\|EPI_ISL_647277\|20 | ···················· | ···················· | ····················· | ···················· | ···················· | ···················· |
| R | 1516 hCoV-19/England/CAMC-B21824/2020\|EPI_ISL_647278\|20 | ···················· | ···················· | ····················· | ···················· | ···················· | ···················· |
| R | 1517 hCoV-19/England/CAMC-B21AE2/2020\|EPI_ISL_647281\|20 | ···················· | ···················· | ····················· | ···················· | ···················· | ···················· |
| R | 1518 hCoV-19/England/CAMC-B21B85/2020\|EPI_ISL_647282\|20 | ···················· | ···················· | ····················· | ···················· | ···················· | ···················· |
| R | 1519 hCoV-19/England/CAMC-B22BEE/2020\|EPI_ISL_647283\|20 | ···················· | ···················· | ····················· | ···················· | ···················· | ···················· |
| R | 1520 hCoV-19/England/CAMC-B2296C/2020\|EPI_ISL_647285\|20 | ···················· | ···················· | ····················· | ···················· | ···················· | ···················· |
| R | 1521 hCoV-19/England/CAMC-B22683/2020\|EPI_ISL_647286\|20 | ···················· | ···················· | ····················· | ···················· | ···················· | ···················· |
| R | 1522 hCoV-19/England/CAMC-B221A0/2020\|EPI_ISL_647287\|20 | ···················· | ···················· | ····················· | ···················· | ···················· | ···················· |
| R | 1523 hCoV-19/England/CAMC-B2146C/2020\|EPI_ISL_647288\|20 | ···················· | ···················· | ····················· | ···················· | ···················· | ···················· |
| R | 1524 hCoV-19/USA/CA-CZB-10299/2020\|EPI_ISL_648500\|2020- | ···················· | ·A·················· | ····················· | ···················· | ···················· | ···················· |

| | | | | | | | |
|---|---|---|---|---|---|---|---|
| R | 1526 hCoV-19/USA/CA-CZB-12383/2020\|EPI_ISL_648505\|2020- | ···················· | ···················· | ···················· | ···················· | ···················· | ···················· |
| R | 1527 hCoV-19/USA/CA-CZB-12384/2020\|EPI_ISL_648506\|2020- | ···················· | ···················· | ···················· | ···················· | ···················· | ···················· |
| R | 1528 hCoV-19/USA/CA-CZB-12387/2020\|EPI_ISL_648508\|2020- | ···················· | ···················· | ···················· | ···················· | ···················· | ···················· |
| R | 1529 hCoV-19/USA/CA-CZB-12390/2020\|EPI_ISL_648509\|2020- | ···················· | ···················· | ···················· | ···················· | ···················· | ···················· |
| R | 1530 hCoV-19/USA/CA-CZB-12392/2020\|EPI_ISL_648510\|2020- | ···················· | ···················· | ···················· | ···················· | ···················· | ···················· |
| R | 1531 hCoV-19/USA/CA-CZB-12393/2020\|EPI_ISL_648511\|2020- | ···················· | ···················· | ···················· | ···················· | ···················· | ···················· |
| R | 1532 hCoV-19/USA/CA-CZB-12394/2020\|EPI_ISL_648512\|2020- | ···················· | ···················· | ···················· | ···················· | ···················· | ···················· |
| R | 1533 hCoV-19/USA/CA-CZB-12857/2020\|EPI_ISL_648513\|2020- | ···················· | ···················· | ···················· | ···················· | ···················· | ···················· |
| R | 1534 hCoV-19/USA/CA-CZB-12858/2020\|EPI_ISL_648514\|2020- | ···················· | ···················· | ···················· | ···················· | ···················· | ···················· |
| R | 1535 hCoV-19/USA/CA-CZB-12859/2020\|EPI_ISL_648515\|2020- | ···················· | ···················· | ···················· | ···················· | ···················· | ···················· |
| R | 1536 hCoV-19/USA/CA-CZB-12860/2020\|EPI_ISL_648516\|2020- | ···················· | ···················· | ···················· | ···················· | ···················· | ···················· |
| R | 1537 hCoV-19/Australia/VIC16215/2020\|EPI_ISL_641050\|202 | ···················· | ···················· | ···················· | ···················· | ···················· | ···················· |
| R | 1538 hCoV-19/Australia/VIC16093/2020\|EPI_ISL_641051\|202 | ···················· | ···················· | ···················· | ···················· | ···················· | ···················· |
| R | 1542 hCoV-19/Australia/VIC16624/2020\|EPI_ISL_641058\|202 | ···················· | ···················· | ···················· | ···················· | ···················· | ···················· |
| R | 1543 hCoV-19/Australia/VIC11619/2020\|EPI_ISL_641060\|202 | ···················· | ···················· | ···················· | ···················· | ···················· | ···················· |
| R | 1544 hCoV-19/Australia/VIC13016/2020\|EPI_ISL_641061\|202 | ···················· | ···················· | ···················· | ···················· | ···················· | ···················· |
| R | 1545 hCoV-19/Australia/VIC12152/2020\|EPI_ISL_641063\|202 | ···················· | ···················· | ···················· | ···················· | ···················· | ···················· |
| R | 1546 hCoV-19/Australia/VIC12918/2020\|EPI_ISL_641064\|202 | ···················· | ···················· | ···················· | ···················· | ···················· | ···················· |
| R | 1547 hCoV-19/Australia/VIC12116/2020\|EPI_ISL_641065\|202 | ···················· | ···················· | ···················· | ···················· | ···················· | ···················· |
| R | 1548 hCoV-19/Australia/VIC12049/2020\|EPI_ISL_641066\|202 | ···················· | ···················· | ···················· | ···················· | ···················· | ···················· |
| R | 1549 hCoV-19/Australia/VIC13192/2020\|EPI_ISL_641081\|202 | ···················· | ···················· | ···················· | ···················· | ···················· | ···················· |
| R | 1550 hCoV-19/Australia/VIC13077/2020\|EPI_ISL_641085\|202 | ···················· | ···················· | ···················· | ···················· | ···················· | ···················· |
| R | 1551 hCoV-19/Australia/VIC12808/2020\|EPI_ISL_641089\|202 | ···················· | ···················· | ···················· | ···················· | ···················· | ···················· |
| R | 1552 hCoV-19/England/QEUH-B11AA9/2020\|EPI_ISL_642380\|20 | ···················· | ···················· | ···················· | ···················· | ···················· | ···················· |
| R | 1553 hCoV-19/England/QEUH-B11BC4/2020\|EPI_ISL_642381\|20 | ···················· | ···················· | ···················· | ···················· | ···················· | ···················· |
| R | 1554 hCoV-19/England/QEUH-B1165A/2020\|EPI_ISL_642383\|20 | ···················· | ···················· | ···················· | ···················· | ···················· | ···················· |
| R | 1555 hCoV-19/England/QEUH-B11B79/2020\|EPI_ISL_642384\|20 | ···················· | ···················· | ···················· | ···················· | ···················· | ···················· |
| R | 1556 hCoV-19/England/QEUH-B11E61/2020\|EPI_ISL_642385\|20 | ···················· | ···················· | ···················· | ···················· | ···················· | ···················· |
| R | 1557 hCoV-19/England/QEUH-B11827/2020\|EPI_ISL_642387\|20 | ···················· | ···················· | ···················· | ···················· | ···················· | ···················· |
| R | 1558 hCoV-19/England/QEUH-B1163C/2020\|EPI_ISL_642388\|20 | ···················· | ·A·················· | ···················· | ···················· | ···················· | ···················· |
| R | 1559 hCoV-19/England/QEUH-B11784/2020\|EPI_ISL_642389\|20 | ···················· | ···················· | ···················· | ···················· | ···················· | ···················· |
| R | 1560 hCoV-19/England/QEUH-B128BD/2020\|EPI_ISL_642390\|20 | ···················· | ···················· | ···················· | ···················· | ···················· | ···················· |
| R | 1561 hCoV-19/England/QEUH-B12756/2020\|EPI_ISL_642391\|20 | ···················· | ···················· | ···················· | ···················· | ···················· | ···················· |
| R | 1562 hCoV-19/England/QEUH-B11DBF/2020\|EPI_ISL_642392\|20 | ···················· | ···················· | ···················· | ···················· | ···················· | ···················· |
| R | 1563 hCoV-19/England/QEUH-B11D37/2020\|EPI_ISL_642393\|20 | ···················· | ···················· | ···················· | ···················· | ···················· | ···················· |
| R | 1564 hCoV-19/England/QEUH-B11CC1/2020\|EPI_ISL_642394\|20 | ···················· | ···················· | ···················· | ···················· | ···················· | ···················· |
| R | 1565 hCoV-19/England/QEUH-B119D9/2020\|EPI_ISL_642395\|20 | ···················· | ···················· | ···················· | ···················· | ···················· | ···················· |
| R | 1566 hCoV-19/England/QEUH-B128CC/2020\|EPI_ISL_642396\|20 | ···················· | ···················· | ···················· | ···················· | ···················· | ···················· |
| R | 1567 hCoV-19/England/QEUH-B11AD6/2020\|EPI_ISL_642397\|20 | ···················· | ···················· | ···················· | ···················· | ···················· | ···················· |
| R | 1568 hCoV-19/England/QEUH-B1312A/2020\|EPI_ISL_642398\|20 | ···················· | ···················· | ···················· | ···················· | ···················· | ···················· |
| R | 1569 hCoV-19/England/QEUH-B11AB8/2020\|EPI_ISL_642399\|20 | ···················· | ···················· | ···················· | ···················· | ···················· | ···················· |
| R | 1570 hCoV-19/England/CAMC-B0BAB1/2020\|EPI_ISL_643600\|20 | ···················· | ··············C····· | ···················· | ···················· | ···················· | ···················· |
| R | 1571 hCoV-19/England/CAMC-B08243/2020\|EPI_ISL_643601\|20 | ···················· | ···················· | ···················· | ···················· | ···················· | ···················· |
| R | 1572 hCoV-19/England/CAMC-B0B18F/2020\|EPI_ISL_643602\|20 | ···················· | ···················· | ···················· | ···················· | ······T············· | ···················· |
| R | 1573 hCoV-19/England/CAMC-B0C90E/2020\|EPI_ISL_643604\|20 | ···················· | ···················· | ···················· | ···················· | ···················· | ···················· |
| R | 1574 hCoV-19/England/CAMC-B0AC07/2020\|EPI_ISL_643605\|20 | ···················· | ···················· | ···················· | ···················· | ···················· | ···················· |
| R | 1575 hCoV-19/England/CAMC-B08498/2020\|EPI_ISL_643606\|20 | ···················· | ·A·················· | ···················· | ···················· | ···················· | ···A················ |
| R | 1576 hCoV-19/England/CAMC-B0ABDD/2020\|EPI_ISL_643607\|20 | ···················· | ···················· | ···················· | ···················· | ···················· | ···················· |
| R | 1577 hCoV-19/England/CAMC-B0CDC6/2020\|EPI_ISL_643608\|20 | ···················· | ···················· | ···················· | ···················· | ···················· | ···················· |
| R | 1578 hCoV-19/England/CAMC-B0AF77/2020\|EPI_ISL_643609\|20 | ···················· | ···················· | ···················· | ···················· | ···················· | ···················· |
| R | 1579 hCoV-19/England/CAMC-B0B240/2020\|EPI_ISL_643610\|20 | ···················· | ···················· | ···················· | ···················· | ···················· | ···················· |
| R | 1580 hCoV-19/England/CAMC-B0AE3E/2020\|EPI_ISL_643611\|20 | ···················· | ···················· | ···················· | ···················· | ···················· | ···················· |
| R | 1581 hCoV-19/England/CAMC-B0AEB6/2020\|EPI_ISL_643612\|20 | ···················· | ···················· | ···················· | ···················· | ···················· | ···················· |
| R | 1582 hCoV-19/England/CAMC-B0AEE3/2020\|EPI_ISL_643613\|20 | ···················· | ···················· | ···················· | ···················· | ···················· | ···················· |
| R | 1583 hCoV-19/England/CAMC-B0CCE7/2020\|EPI_ISL_643614\|20 | ···················· | ···················· | ···················· | ···················· | ···················· | ···················· |
| R | 1584 hCoV-19/England/CAMC-B0AAC1/2020\|EPI_ISL_643615\|20 | ···················· | ···················· | ···················· | ···················· | ···················· | ···················· |
| R | 1585 hCoV-19/England/CAMC-B0B2C8/2020\|EPI_ISL_643616\|20 | ···················· | ···················· | ···················· | ···················· | ···················· | ···················· |
| R | 1586 hCoV-19/England/CAMC-B0846B/2020\|EPI_ISL_643617\|20 | ···················· | ···················· | ···················· | ···················· | ···················· | ···················· |
| R | 1587 hCoV-19/England/CAMC-B0CB9F/2020\|EPI_ISL_643618\|20 | ···················· | ···················· | ···················· | ···················· | ···················· | ···················· |
| R | 1588 hCoV-19/England/CAMC-B0ADB9/2020\|EPI_ISL_643619\|20 | ···················· | ···················· | ···················· | ···················· | ···················· | ···················· |
| R | 1589 hCoV-19/England/CAMC-B0B3B6/2020\|EPI_ISL_643620\|20 | ···················· | ···················· | ···················· | ···················· | ···················· | ···················· |
| R | 1590 hCoV-19/England/CAMC-B0AC16/2020\|EPI_ISL_643621\|20 | ···················· | ···················· | ···················· | ···················· | ···················· | ···················· |
| R | 1591 hCoV-19/England/CAMC-B0C643/2020\|EPI_ISL_643622\|20 | ···················· | ···················· | ···················· | ···················· | ···················· | ···················· |
| R | 1592 hCoV-19/USA/VA-DCLS-1995/2020\|EPI_ISL_644900\|2020- | ···················· | ···················· | ···················· | ···················· | ···················· | ···················· |
| R | 1593 hCoV-19/USA/VA-DCLS-1996/2020\|EPI_ISL_644901\|2020- | ···················· | ···················· | ···················· | ···················· | ···················· | ···················· |
| R | 1594 hCoV-19/USA/VA-DCLS-2000/2020\|EPI_ISL_644903\|2020- | ···················· | ·A·················· | ···················· | ···················· | ···················· | ···················· |
| R | 1595 hCoV-19/USA/VA-DCLS-2002/2020\|EPI_ISL_644904\|2020- | ···················· | ···················· | ···················· | ···················· | ···················· | ···················· |
| R | 1596 hCoV-19/USA/VA-DCLS-2003/2020\|EPI_ISL_644905\|2020- | ···················· | ···················· | ···················· | ···················· | ···················· | ···················· |
| R | 1597 hCoV-19/USA/VA-DCLS-2005/2020\|EPI_ISL_644906\|2020- | ···················· | ···················· | ···················· | ···················· | ···················· | ···················· |
| R | 1598 hCoV-19/USA/VA-DCLS-2006/2020\|EPI_ISL_644907\|2020- | ···················· | ···················· | ···················· | ···················· | ···················· | ···················· |
| R | 1599 hCoV-19/USA/VA-DCLS-2007/2020\|EPI_ISL_644908\|2020- | ···················· | ···················· | ···················· | ···················· | ···················· | ···················· |
| R | 1600 hCoV-19/USA/VA-DCLS-2008/2020\|EPI_ISL_644909\|2020- | ···················· | ···················· | ···················· | ···················· | ···················· | ···················· |
| R | 1601 hCoV-19/USA/VA-DCLS-2009/2020\|EPI_ISL_644910\|2020- | ···················· | ·A·················· | ···················· | ···················· | ···················· | ···················· |
| R | 1602 hCoV-19/USA/VA-DCLS-1871/2020\|EPI_ISL_644911\|2020- | ···················· | ····A··············· | ···················· | ···················· | ···················· | ···················· |
| R | 1603 hCoV-19/USA/VA-DCLS-1872/2020\|EPI_ISL_644912\|2020- | ···················· | ···················· | ···················· | ···················· | ···················· | ···················· |
| R | 1604 hCoV-19/USA/VA-DCLS-1873/2020\|EPI_ISL_644913\|2020- | ···················· | ·A·················· | ···················· | ···················· | ···················· | ···················· |
| R | 1605 hCoV-19/USA/VA-DCLS-1874/2020\|EPI_ISL_644914\|2020- | ···················· | ·A·················· | ···················· | ···················· | ···················· | ···················· |
| R | 1606 hCoV-19/USA/VA-DCLS-1877/2020\|EPI_ISL_644915\|2020- | ···················· | ·A·················· | ···················· | ···················· | ···················· | ···················· |
| R | 1607 hCoV-19/USA/VA-DCLS-1880/2020\|EPI_ISL_644916\|2020- | ···················· | ···················· | ···················· | ···················· | ···················· | ···················· |
| R | 1608 hCoV-19/USA/VA-DCLS-1883/2020\|EPI_ISL_644917\|2020- | ···················· | ···················· | ···················· | ···················· | ···················· | ···················· |

| | | | | | | | | |
|---|---|---|---|---|---|---|---|---|
| R | 1609 hCoV-19/USA/VA-DCLS-1886/2020\|EPI_ISL_644919\|2020- | ···················· | ···················· | ···················· | ···················· | ···················· | ···················· | |
| R | 1610 hCoV-19/USA/VA-DCLS-1887/2020\|EPI_ISL_644920\|2020- | ···················· | ···················· | ···················· | ···················· | ···················· | ···················· | |
| R | 1611 hCoV-19/USA/VA-DCLS-1927/2020\|EPI_ISL_644921\|2020- | ···················· | ···················· | ···················· | ···················· | ···················· | ···················· | |
| R | 1612 hCoV-19/USA/VA-DCLS-1928/2020\|EPI_ISL_644922\|2020- | ···················· | ·A·················· | ···················· | ···················· | ···················· | ···················· | |
| R | 1613 hCoV-19/USA/VA-DCLS-1932/2020\|EPI_ISL_644923\|2020- | ···················· | ·A·················· | ···················· | ···················· | ···················· | ···················· | |
| R | 1614 hCoV-19/USA/VA-DCLS-1933/2020\|EPI_ISL_644924\|2020- | ···················· | ···················· | ···················· | ···················· | ···················· | ···················· | |
| R | 1615 hCoV-19/USA/VA-DCLS-1935/2020\|EPI_ISL_644925\|2020- | ···················· | ···················· | ···················· | ···················· | ···················· | ···················· | |
| R | 1616 hCoV-19/USA/VA-DCLS-1936/2020\|EPI_ISL_644926\|2020- | ···················· | ···················· | ···················· | ···················· | ···················· | ···················· | |
| R | 1617 hCoV-19/USA/VA-DCLS-1939/2020\|EPI_ISL_644927\|2020- | ···················· | ···················· | ···················· | ···················· | ···················· | ···················· | |
| R | 1618 hCoV-19/USA/VA-DCLS-1941/2020\|EPI_ISL_644928\|2020- | ···················· | ···················· | ···················· | ···················· | ···················· | ···················· | |
| R | 1619 hCoV-19/USA/VA-DCLS-1948/2020\|EPI_ISL_644930\|2020- | ···················· | ·A·················· | ··········T········· | ···················· | ···················· | ···················· | |
| R | 1620 hCoV-19/USA/VA-DCLS-1949/2020\|EPI_ISL_644931\|2020- | ···················· | ·A·················· | ···················· | ···················· | ···················· | ···················· | |
| R | 1621 hCoV-19/USA/VA-DCLS-1951/2020\|EPI_ISL_644932\|2020- | ···················· | ···················· | ···················· | ···················· | ···················· | ···················· | |
| R | 1622 hCoV-19/USA/MN-MDH-2002/2020\|EPI_ISL_644933\|2020-1 | ···················· | ·A·················· | ···················· | ···················· | ···················· | ···················· | |
| R | 1623 hCoV-19/USA/MN-MDH-2003/2020\|EPI_ISL_644934\|2020-1 | ···················· | ·A·················· | ···················· | ···················· | ···················· | ···················· | |
| R | 1624 hCoV-19/USA/MN-MDH-2004/2020\|EPI_ISL_644935\|2020-1 | ···················· | ·A·················· | ···················· | ···················· | ···················· | ···················· | |
| R | 1625 hCoV-19/USA/MN-MDH-2005/2020\|EPI_ISL_644936\|2020-1 | ···················· | ·A·················· | ···················· | ···················· | NNN················· | ···················· | 1 |
| R | 1626 hCoV-19/USA/MN-MDH-2006/2020\|EPI_ISL_644937\|2020-0 | ···················· | ·A·················· | ···················· | ···················· | ···················· | ···················· | |
| R | 1627 hCoV-19/USA/MN-MDH-2007/2020\|EPI_ISL_644938\|2020-0 | ···················· | ···················· | ···················· | ···················· | ···················· | ···················· | |
| R | 1628 hCoV-19/USA/MN-MDH-2008/2020\|EPI_ISL_644939\|2020-0 | ···················· | ·A·················· | ···················· | ···················· | ···················· | ···················· | |
| R | 1629 hCoV-19/USA/MN-MDH-2009/2020\|EPI_ISL_644940\|2020-0 | ···················· | ···················· | ···················· | ···················· | ···················· | ···················· | |
| R | 1630 hCoV-19/USA/MN-MDH-2010/2020\|EPI_ISL_644941\|2020-1 | ···················· | ···················· | ···················· | ···················· | ···················· | ···················· | |
| R | 1631 hCoV-19/USA/MN-MDH-2011/2020\|EPI_ISL_644942\|2020-1 | ···················· | ···················· | ···················· | ···················· | ···················· | ···················· | |
| R | 1632 hCoV-19/USA/MN-MDH-2012/2020\|EPI_ISL_644943\|2020-1 | ···················· | ···················· | ···················· | ···················· | ···················· | ···················· | |
| R | 1633 hCoV-19/USA/MN-MDH-2013/2020\|EPI_ISL_644944\|2020-0 | ···················· | ···················· | ···················· | ···················· | ···················· | ···················· | |
| R | 1634 hCoV-19/Japan/Donner39/2020\|EPI_ISL_644945\|2020-05 | ···················· | ···················· | ···················· | ···················· | ···················· | ···················· | |
| R | 1635 hCoV-19/Japan/Donner41/2020\|EPI_ISL_644946\|2020-06 | ···················· | ···················· | ···················· | ···················· | ···················· | ···················· | |
| R | 1636 hCoV-19/Japan/Donner42/2020\|EPI_ISL_644947\|2020-06 | ···················· | ···················· | ···················· | ···················· | ···················· | ···················· | |
| R | 1637 hCoV-19/Japan/Donner43/2020\|EPI_ISL_644948\|2020-06 | ···················· | ···················· | ···················· | ···················· | ···················· | ···················· | |
| R | 1638 hCoV-19/Japan/Donner44/2020\|EPI_ISL_644949\|2020-06 | ···················· | ···················· | ···················· | ···················· | ···················· | ···················· | |
| R | 1639 hCoV-19/Japan/Donner45/2020\|EPI_ISL_644950\|2020-06 | ···················· | ···················· | ···················· | ···················· | ···················· | ···················· | |
| R | 1640 hCoV-19/Japan/Donner46/2020\|EPI_ISL_644951\|2020-06 | ···················· | ···················· | ···················· | ···················· | ···················· | ···················· | |
| R | 1641 hCoV-19/Japan/Donner47/2020\|EPI_ISL_644952\|2020-06 | ···················· | ···················· | ···················· | ···················· | ···················· | ···················· | |
| R | 1642 hCoV-19/Japan/Donner48/2020\|EPI_ISL_644953\|2020-06 | ···················· | ···················· | ···················· | ···················· | ···················· | ···················· | |
| R | 1643 hCoV-19/England/ALDP-B24B37/2020\|EPI_ISL_647100\|20 | ···················· | ·A·················· | ···················· | ···················· | ···················· | ··A················· | |
| R | 1644 hCoV-19/England/ALDP-B23998/2020\|EPI_ISL_647101\|20 | ···················· | ···················· | ···················· | ···················· | ···················· | ···················· | |
| R | 1645 hCoV-19/England/QEUH-B2DA9B/2020\|EPI_ISL_647102\|20 | ···················· | ···················· | ···················· | ···················· | ···················· | ···················· | |
| R | 1646 hCoV-19/England/ALDP-B245EE/2020\|EPI_ISL_647104\|20 | ···················· | ···················· | ···················· | ···················· | ···················· | ···················· | |
| R | 1647 hCoV-19/England/QEUH-B2D9E9/2020\|EPI_ISL_647105\|20 | ···················· | ···················· | ···················· | ···················· | ···················· | ···················· | |
| R | 1648 hCoV-19/England/ALDP-B249D3/2020\|EPI_ISL_647106\|20 | ···················· | ···················· | ···················· | ···················· | ···················· | ···················· | |
| R | 1649 hCoV-19/England/ALDP-B24CAD/2020\|EPI_ISL_647107\|20 | ···················· | ···················· | ···················· | ···················· | ···················· | ···················· | |
| R | 1650 hCoV-19/England/ALDP-B2443C/2020\|EPI_ISL_647108\|20 | ···················· | ···················· | ···················· | ···················· | ···················· | ···················· | |
| R | 1651 hCoV-19/England/QEUH-B2D785/2020\|EPI_ISL_647109\|20 | ···················· | ···················· | ···················· | ···················· | ·····T·············· | ···················· | |
| R | 1652 hCoV-19/England/QEUH-B2ED28/2020\|EPI_ISL_647110\|20 | ···················· | ···················· | ···················· | ···················· | ···················· | ···················· | |
| R | 1653 hCoV-19/England/ALDP-B23673/2020\|EPI_ISL_647111\|20 | ···················· | ···················· | ···················· | ···················· | ···················· | ···················· | |
| R | 1654 hCoV-19/England/ALDP-B24AD0/2020\|EPI_ISL_647112\|20 | ···················· | ···················· | ···················· | ···················· | ···················· | ···················· | |
| R | 1655 hCoV-19/England/ALDP-B237AD/2020\|EPI_ISL_647113\|20 | ···················· | ···················· | ···················· | ···················· | ···················· | ···················· | |
| R | 1656 hCoV-19/England/QEUH-B2D794/2020\|EPI_ISL_647114\|20 | ···················· | ···················· | ···················· | ···················· | ···················· | ···················· | |
| R | 1657 hCoV-19/England/QEUH-B2D749/2020\|EPI_ISL_647115\|20 | ···················· | ···················· | ···················· | ···················· | ···················· | ···················· | |
| R | 1658 hCoV-19/England/ALDP-B245FD/2020\|EPI_ISL_647116\|20 | ···················· | ···················· | ···················· | ···················· | ···················· | ···················· | |
| R | 1659 hCoV-19/England/QEUH-B2DC95/2020\|EPI_ISL_647117\|20 | ···················· | ···················· | ···················· | ···················· | ···················· | ···················· | |
| R | 1660 hCoV-19/England/ALDP-B23A2C/2020\|EPI_ISL_647118\|20 | ···················· | ···················· | ···················· | ···················· | ···················· | ···················· | |
| R | 1661 hCoV-19/England/ALDP-B24250/2020\|EPI_ISL_647119\|20 | ···················· | ···················· | ···················· | ···················· | ···················· | ···················· | |
| R | 1662 hCoV-19/England/ALDP-B23989/2020\|EPI_ISL_647120\|20 | ···················· | ···················· | ···················· | ···················· | ···················· | ···················· | |
| R | 1663 hCoV-19/England/ALDP-B239B6/2020\|EPI_ISL_647122\|20 | ···················· | ···················· | ···················· | ···················· | ···················· | ···················· | |
| R | 1664 hCoV-19/England/CAMC-B22498/2020\|EPI_ISL_647123\|20 | ···················· | ···················· | ···················· | ···················· | ···················· | ···················· | |
| R | 1665 hCoV-19/England/CAMC-B22717/2020\|EPI_ISL_647124\|20 | ···················· | ···················· | ···················· | ···················· | ···················· | ···················· | |
| R | 1666 hCoV-19/England/CAMC-B22E03/2020\|EPI_ISL_647125\|20 | ···················· | ···················· | ···················· | ···················· | ···················· | ···················· | |
| R | 1667 hCoV-19/England/CAMC-B20FF3/2020\|EPI_ISL_647126\|20 | ···················· | ···················· | ···················· | ···················· | ···················· | ···················· | |
| R | 1668 hCoV-19/England/CAMC-B214E4/2020\|EPI_ISL_647127\|20 | ···················· | ···················· | ···················· | ···················· | ···················· | ···················· | |
| R | 1669 hCoV-19/England/CAMC-B22CFA/2020\|EPI_ISL_647129\|20 | ···················· | ···················· | ···················· | ···················· | ···················· | ···················· | |
| R | 1670 hCoV-19/England/CAMC-B22559/2020\|EPI_ISL_647131\|20 | ···················· | ···················· | ···················· | ···················· | ···················· | ···················· | |
| R | 1671 hCoV-19/England/CAMC-B218D9/2020\|EPI_ISL_647134\|20 | ···················· | ···················· | ···················· | ···················· | ···················· | ···················· | |
| R | 1672 hCoV-19/England/CAMC-B2188E/2020\|EPI_ISL_647135\|20 | ···················· | ·A·················· | ···················· | ···················· | ···················· | ··A················· | |
| R | 1673 hCoV-19/England/CAMC-B22C27/2020\|EPI_ISL_647136\|20 | ···················· | ···················· | ···················· | ···················· | ···················· | ···················· | |
| R | 1674 hCoV-19/England/CAMC-B217DC/2020\|EPI_ISL_647137\|20 | ···················· | ···················· | ···················· | ···················· | ·····T·············· | ···················· | |
| R | 1675 hCoV-19/England/CAMC-B21745/2020\|EPI_ISL_647138\|20 | ···················· | ···················· | ···················· | ···················· | ·····T·············· | ···················· | |
| R | 1676 hCoV-19/England/CAMC-B22902/2020\|EPI_ISL_647139\|20 | ···················· | ···················· | ···················· | ···················· | ···················· | ···················· | |
| R | 1677 hCoV-19/England/CAMC-B22F6A/2020\|EPI_ISL_647141\|20 | ···················· | ···················· | ···················· | ···················· | ···················· | ···················· | |
| R | 1678 hCoV-19/England/CAMC-B22C18/2020\|EPI_ISL_647142\|20 | ···················· | ···················· | ···················· | ···················· | ···················· | ···················· | |
| R | 1679 hCoV-19/England/CAMC-B22AD2/2020\|EPI_ISL_647143\|20 | ···················· | ···················· | ···················· | ···················· | ···················· | ···················· | |
| R | 1680 hCoV-19/England/CAMC-B21F5C/2020\|EPI_ISL_647144\|20 | ···················· | ···················· | ···················· | ···················· | ···················· | ···················· | |
| R | 1681 hCoV-19/England/CAMC-B21BC1/2020\|EPI_ISL_647145\|20 | ···················· | ···················· | ···················· | ···················· | ···················· | ···················· | |
| R | 1682 hCoV-19/England/CAMC-B219B8/2020\|EPI_ISL_647146\|20 | ···················· | ···················· | ···················· | ···················· | ···················· | ···················· | |
| R | 1683 hCoV-19/England/CAMC-B22850/2020\|EPI_ISL_647147\|20 | ···················· | ···················· | ···················· | ···················· | ···················· | ···················· | |
| R | 1684 hCoV-19/England/CAMC-B21EB9/2020\|EPI_ISL_647149\|20 | ···················· | ···················· | ···················· | ···················· | ···················· | ···················· | |
| R | 1685 hCoV-19/England/CAMC-B2244D/2020\|EPI_ISL_647150\|20 | ···················· | ···················· | ···················· | ···················· | ···················· | ···················· | |
| R | 1686 hCoV-19/England/CAMC-B21639/2020\|EPI_ISL_647151\|20 | ···················· | ···················· | ···················· | ···················· | ···················· | ···················· | |
| R | 1687 hCoV-19/England/CAMC-B218CA/2020\|EPI_ISL_647153\|20 | ···················· | ···················· | ···················· | ···················· | ···················· | ···················· | |
| R | 1688 hCoV-19/England/CAMC-B20FC6/2020\|EPI_ISL_647154\|20 | ···················· | ···················· | ···················· | ···················· | ···················· | ···················· | |

| | | | | | | | | | |
|---|---|---|---|---|---|---|---|---|---|
| R | 1689 | hCoV-19/England/CAMC-B21C0A/2020\|EPI_ISL_647155\|20 | ···················· | ···················· | ···················· | ···················· | ···················· | ···················· | |
| R | 1690 | hCoV-19/England/CAMC-B2144E/2020\|EPI_ISL_647156\|20 | ···················· | ···················· | ···················· | ···················· | ···················· | ···················· | |
| R | 1691 | hCoV-19/England/CAMC-B219D6/2020\|EPI_ISL_647157\|20 | ···················· | ···················· | ···················· | ···················· | ···················· | ···················· | |
| R | 1692 | hCoV-19/England/CAMC-B21833/2020\|EPI_ISL_647158\|20 | ···················· | ···················· | ···················· | ···················· | ···················· | ···················· | |
| R | 1693 | hCoV-19/England/CAMC-B22A78/2020\|EPI_ISL_647159\|20 | ···················· | ···················· | ···················· | ···················· | ···················· | ···················· | |
| R | 1694 | hCoV-19/England/CAMC-B2253B/2020\|EPI_ISL_647164\|20 | ···················· | ···················· | ···················· | ···················· | ······T············· | ···················· | |
| R | 1695 | hCoV-19/England/CAMC-B22CAF/2020\|EPI_ISL_647166\|20 | ···················· | ···················· | ···················· | ···················· | ······T············· | ···················· | |
| R | 1696 | hCoV-19/England/CAMC-B22629/2020\|EPI_ISL_647167\|20 | ···················· | ···················· | ···················· | ···················· | ···················· | ···················· | |
| R | 1697 | hCoV-19/England/CAMC-B22E7C/2020\|EPI_ISL_647170\|20 | ···················· | ···················· | ···················· | ···················· | ···················· | ···················· | |
| R | 1698 | hCoV-19/England/CAMC-B224B6/2020\|EPI_ISL_647171\|20 | ···················· | ···················· | ···················· | ···················· | ···················· | ···················· | |
| R | 1699 | hCoV-19/England/CAMC-B21DBC/2020\|EPI_ISL_647172\|20 | ···················· | ···················· | ···················· | ···················· | ···················· | ···················· | |
| R | 1700 | hCoV-19/England/CAMC-B22BFD/2020\|EPI_ISL_647173\|20 | ···················· | ···················· | ···················· | ···················· | ···················· | ···················· | |
| R | 1701 | hCoV-19/England/CAMC-B22CBE/2020\|EPI_ISL_647174\|20 | ···················· | ···················· | ···················· | ···················· | ····T··············· | ···················· | |
| R | 1702 | hCoV-19/England/CAMC-B22CEB/2020\|EPI_ISL_647176\|20 | ···················· | ···················· | ···················· | ···················· | ···················· | ···················· | |
| R | 1703 | hCoV-19/England/CAMC-B215F0/2020\|EPI_ISL_647177\|20 | ···················· | ···················· | ···················· | ···················· | ······T············· | ···················· | |
| R | 1704 | hCoV-19/England/CAMC-B21DE9/2020\|EPI_ISL_647178\|20 | ···················· | ···················· | ···················· | ···················· | ···················· | ···················· | |
| R | 1705 | hCoV-19/England/CAMC-B2161B/2020\|EPI_ISL_647179\|20 | ···················· | ···················· | ···················· | ···················· | ···················· | ···················· | |
| R | 1706 | hCoV-19/England/CAMC-B2289C/2020\|EPI_ISL_647180\|20 | ···················· | ···················· | ···················· | ···················· | ···················· | ···················· | |
| R | 1707 | hCoV-19/England/CAMC-B232E8/2020\|EPI_ISL_647181\|20 | ···················· | ···················· | ···················· | ···················· | ···················· | ···················· | |
| R | 1708 | hCoV-19/England/CAMC-B226B0/2020\|EPI_ISL_647183\|20 | ···················· | ···················· | ···················· | ···················· | ···················· | ···················· | |
| R | 1709 | hCoV-19/England/CAMC-B22674/2020\|EPI_ISL_647184\|20 | ···················· | ···················· | ···················· | ···················· | ···················· | ···················· | |
| R | 1710 | hCoV-19/England/CAMC-B21709/2020\|EPI_ISL_647185\|20 | ···················· | ···················· | ···················· | ···················· | ···················· | ···················· | |
| R | 1711 | hCoV-19/England/CAMC-B22B0C/2020\|EPI_ISL_647186\|20 | ···················· | ·A·················· | ···················· | ···················· | ···················· | ···················· | |
| R | 1712 | hCoV-19/England/CAMC-B2189D/2020\|EPI_ISL_647187\|20 | ···················· | ·A·················· | ···················· | ···················· | ···················· | ···················· | |
| R | 1713 | hCoV-19/England/CAMC-B215D2/2020\|EPI_ISL_647189\|20 | ···················· | ···················· | ···················· | ···················· | ···················· | ···················· | |
| R | 1714 | hCoV-19/England/CAMC-B22AE1/2020\|EPI_ISL_647191\|20 | ···················· | ···················· | ···················· | ···················· | ···················· | ···················· | |
| R | 1715 | hCoV-19/England/CAMC-B22AA5/2020\|EPI_ISL_647192\|20 | ···················· | ···················· | ···················· | ···················· | ······T············· | ···················· | |
| R | 1716 | hCoV-19/England/CAMC-B22F4C/2020\|EPI_ISL_647193\|20 | ···················· | ···················· | ···················· | ···················· | ······T············· | ···················· | |
| R | 1717 | hCoV-19/England/CAMC-B20FB7/2020\|EPI_ISL_647194\|20 | ···················· | ···················· | ···················· | ···················· | ······T············· | ···················· | |
| R | 1718 | hCoV-19/England/CAMC-B21675/2020\|EPI_ISL_647195\|20 | ···················· | ···················· | ···················· | ···················· | ···················· | ···················· | |
| R | 1719 | hCoV-19/England/CAMC-B21B2B/2020\|EPI_ISL_647197\|20 | ···················· | ···················· | ···················· | ···················· | ···················· | ···················· | |
| R | 1720 | hCoV-19/England/CAMC-B228BA/2020\|EPI_ISL_647199\|20 | ···················· | ···················· | ···················· | ···················· | ···················· | ···················· | |
| R | 1721 | hCoV-19/USA/CA-CZB-12319/2020\|EPI_ISL_648401\|2020- | ···················· | ···················· | ···················· | ···················· | ···················· | ···················· | |
| R | 1722 | hCoV-19/USA/CA-CZB-12322/2020\|EPI_ISL_648403\|2020- | ···················· | ···················· | ···················· | ···················· | ···················· | ···················· | |
| R | 1723 | hCoV-19/USA/CA-CZB-12329/2020\|EPI_ISL_648407\|2020- | ···················· | ·A·················· | ···················· | ···················· | ···················· | ···················· | |
| R | 1724 | hCoV-19/USA/CA-CZB-12330/2020\|EPI_ISL_648408\|2020- | ···················· | ·A·················· | ···················· | ···················· | ···················· | ···················· | |
| R | 1725 | hCoV-19/USA/CA-CZB-12332/2020\|EPI_ISL_648409\|2020- | ···················· | ·A·················· | ···················· | ···················· | N··················· | ···················· | |
| R | 1728 | hCoV-19/USA/CA-CZB-12335/2020\|EPI_ISL_648412\|2020- | ···················· | ·A·················· | ···················· | ···················· | ···················· | ···················· | |
| R | 1729 | hCoV-19/USA/CA-CZB-12340/2020\|EPI_ISL_648415\|2020- | ···················· | ···················· | ···················· | ···················· | ···················· | ···················· | |
| R | 1730 | hCoV-19/USA/CA-CZB-12341/2020\|EPI_ISL_648416\|2020- | ···················· | ···················· | ···················· | ···················· | ···················· | ···················· | |
| R | 1732 | hCoV-19/USA/CA-CZB-12345/2020\|EPI_ISL_648419\|2020- | ···················· | ·A·················· | ···················· | ···················· | ···················· | ···················· | |
| R | 1733 | hCoV-19/USA/CA-CZB-12346/2020\|EPI_ISL_648420\|2020- | ···················· | ·A·················· | ···················· | ···················· | ···················· | ···················· | |
| R | 1734 | hCoV-19/USA/CA-CZB-12348/2020\|EPI_ISL_648421\|2020- | ···················· | ·A·················· | ···················· | ···················· | ···················· | ···················· | |
| R | 1735 | hCoV-19/USA/CA-CZB-12351/2020\|EPI_ISL_648423\|2020- | ···················· | ···················· | ···················· | ···················· | ···················· | ···················· | |
| R | 1736 | hCoV-19/USA/CA-CZB-12352/2020\|EPI_ISL_648424\|2020- | ···················· | ·············A······ | ···················· | ···················· | ···················· | ···················· | |
| R | 1737 | hCoV-19/USA/CA-CZB-12353/2020\|EPI_ISL_648425\|2020- | ···················· | ·A·················· | ···················· | ···················· | ···················· | ···················· | |
| R | 1738 | hCoV-19/USA/CA-CZB-12354/2020\|EPI_ISL_648426\|2020- | ···················· | ·A·················· | ···················· | ···················· | ···················· | ···················· | |
| R | 1739 | hCoV-19/USA/CA-CZB-12355/2020\|EPI_ISL_648427\|2020- | ···················· | ·A·················· | ···················· | ···················· | ···················· | ···················· | |
| R | 1740 | hCoV-19/USA/CA-CZB-12356/2020\|EPI_ISL_648428\|2020- | ···················· | ·A·················· | ···················· | ···················· | ···················· | ···················· | |
| R | 1741 | hCoV-19/USA/CA-CZB-12358/2020\|EPI_ISL_648429\|2020- | ···················· | ·A·················· | ···················· | ···················· | ···················· | ···················· | |
| R | 1742 | hCoV-19/USA/CA-CZB-12359/2020\|EPI_ISL_648430\|2020- | ···················· | ·A·················· | ···················· | ···················· | ···················· | ···················· | |
| R | 1743 | hCoV-19/USA/CA-CZB-12360/2020\|EPI_ISL_648431\|2020- | ···················· | ·A·················· | ···················· | ···················· | ···················· | ···················· | |
| R | 1744 | hCoV-19/USA/CA-CZB-12362/2020\|EPI_ISL_648432\|2020- | ···················· | ·A·················· | ···················· | ···················· | ···················· | ···················· | |
| R | 1745 | hCoV-19/USA/CA-CZB-12363/2020\|EPI_ISL_648433\|2020- | ···················· | ·A·················· | ···················· | ···················· | ···················· | ···················· | |
| R | 1746 | hCoV-19/USA/CA-CZB-12364/2020\|EPI_ISL_648434\|2020- | ···················· | ·A·················· | ···················· | ···················· | ···················· | ···················· | |
| R | 1747 | hCoV-19/USA/CA-CZB-12365/2020\|EPI_ISL_648435\|2020- | ···················· | ·A·················· | ···················· | ···················· | ···················· | ···················· | |
| R | 1748 | hCoV-19/USA/CA-CZB-12366/2020\|EPI_ISL_648436\|2020- | ···················· | ·A·················· | ···················· | ···················· | ···················· | ···················· | |
| R | 1749 | hCoV-19/USA/CA-CZB-12774/2020\|EPI_ISL_648437\|2020- | ···················· | ·A·················· | ···················· | ···················· | ···················· | ···················· | |
| R | 1750 | hCoV-19/USA/CA-CZB-12775/2020\|EPI_ISL_648438\|2020- | ···················· | ·A·················· | ···················· | ···················· | ···················· | ···················· | |
| R | 1751 | hCoV-19/USA/CA-CZB-12776/2020\|EPI_ISL_648439\|2020- | ···················· | ···················· | ···················· | ···················· | ···················· | ···················· | |
| R | 1752 | hCoV-19/USA/CA-CZB-12777/2020\|EPI_ISL_648440\|2020- | ···················· | ···················· | ···················· | ···················· | ···················· | ···················· | |
| R | 1753 | hCoV-19/USA/CA-CZB-12779/2020\|EPI_ISL_648441\|2020- | ···················· | ·A·················· | ···················· | ···················· | ···················· | ···················· | |
| R | 1754 | hCoV-19/USA/CA-CZB-12786/2020\|EPI_ISL_648443\|2020- | ···················· | ·A·················· | ···················· | ···················· | ···················· | ···················· | |
| R | 1755 | hCoV-19/USA/CA-CZB-12787/2020\|EPI_ISL_648444\|2020- | ···················· | ·A·················· | ···················· | ···················· | ···················· | ···················· | |
| R | 1756 | hCoV-19/USA/CA-CZB-12788/2020\|EPI_ISL_648445\|2020- | ···················· | ···················· | ···················· | ···················· | ···················· | ···················· | |
| R | 1757 | hCoV-19/USA/CA-CZB-12791/2020\|EPI_ISL_648447\|2020- | ···················· | ·A·················· | ···················· | ···················· | ···················· | ···················· | |
| R | 1758 | hCoV-19/USA/CA-CZB-12792/2020\|EPI_ISL_648448\|2020- | ···················· | ·A·················· | ···················· | ···················· | ···················· | ···················· | |
| R | 1759 | hCoV-19/USA/CA-CZB-12793/2020\|EPI_ISL_648449\|2020- | ···················· | ·A·················· | ···················· | ···················· | ···················· | ···················· | |
| R | 1760 | hCoV-19/USA/CA-CZB-12795/2020\|EPI_ISL_648450\|2020- | ···················· | ···················· | ···················· | ···················· | ···················· | ···················· | |
| R | 1762 | hCoV-19/USA/CA-CZB-12798/2020\|EPI_ISL_648452\|2020- | ···················· | ·A·················· | ···················· | ···················· | ···················· | ···················· | |
| R | 1764 | hCoV-19/USA/CA-CZB-12802/2020\|EPI_ISL_648454\|2020- | ···················· | ·A·················· | ···················· | ···················· | ···················· | ···················· | |
| R | 1765 | hCoV-19/USA/CA-CZB-12804/2020\|EPI_ISL_648455\|2020- | ···················· | ·A·················· | ···················· | ···················· | ···················· | ···················· | |
| R | 1766 | hCoV-19/USA/CA-CZB-12805/2020\|EPI_ISL_648456\|2020- | ···················· | ·A·················· | ···················· | ···················· | ···················· | ···················· | |
| R | 1767 | hCoV-19/USA/CA-CZB-12808/2020\|EPI_ISL_648459\|2020- | ···················· | ·A·················· | ···················· | ···················· | ···················· | ···················· | |
| R | 1770 | hCoV-19/USA/CA-CZB-12811/2020\|EPI_ISL_648462\|2020- | ···················· | ·A·················· | ···················· | ···················· | ···················· | ···················· | |
| R | 1771 | hCoV-19/USA/CA-CZB-12812/2020\|EPI_ISL_648463\|2020- | ···················· | ···················· | ···················· | ···················· | ···················· | ···················· | |
| R | 1772 | hCoV-19/USA/CA-CZB-12814/2020\|EPI_ISL_648465\|2020- | ···················· | ···················· | ···················· | ···················· | ···················· | ···················· | |
| R | 1773 | hCoV-19/USA/CA-CZB-12816/2020\|EPI_ISL_648467\|2020- | ···················· | ···················· | ···················· | ···················· | NNN················· | ···················· | 1 |
| R | 1774 | hCoV-19/USA/CA-CZB-12817/2020\|EPI_ISL_648468\|2020- | ···················· | ···················· | ···················· | ···················· | ···················· | ···················· | |
| R | 1775 | hCoV-19/USA/CA-CZB-12818/2020\|EPI_ISL_648469\|2020- | ···················· | ···················· | ···················· | ···················· | ···················· | ···················· | |

| | | | | | | | |
|---|---|---|---|---|---|---|---|
| R | 1776 hCoV-19/USA/CA-CZB-12819/2020\|EPI_ISL_648470\|2020- | ···················· | ···················· | ····················· | ···················· | ···················· | ···················· |
| R | 1777 hCoV-19/USA/CA-CZB-12820/2020\|EPI_ISL_648471\|2020- | ···················· | ···················· | ····················· | ···················· | ···················· | ···················· |
| R | 1778 hCoV-19/USA/CA-CZB-12821/2020\|EPI_ISL_648472\|2020- | ···················· | ···················· | ····················· | ···················· | ···················· | ···················· |
| R | 1779 hCoV-19/USA/CA-CZB-12822/2020\|EPI_ISL_648473\|2020- | ···················· | ···················· | ····················· | ···················· | ···················· | ···················· |
| R | 1780 hCoV-19/USA/CA-CZB-12823/2020\|EPI_ISL_648474\|2020- | ···················· | ···················· | ····················· | ···················· | ···················· | ···················· |
| R | 1781 hCoV-19/USA/CA-CZB-12824/2020\|EPI_ISL_648475\|2020- | ···················· | ···················· | ····················· | ···················· | ···················· | ···················· |
| R | 1782 hCoV-19/USA/CA-CZB-12825/2020\|EPI_ISL_648476\|2020- | ···················· | ·A·················· | ····················· | ···················· | ···················· | ···················· |
| R | 1783 hCoV-19/USA/CA-CZB-12835/2020\|EPI_ISL_648481\|2020- | ···················· | ·A·················· | ····················· | ···················· | ···················· | ···················· |
| R | 1784 hCoV-19/USA/CA-CZB-12836/2020\|EPI_ISL_648482\|2020- | ···················· | ·A·················· | ····················· | ···················· | ···················· | ···················· |
| R | 1785 hCoV-19/USA/CA-CZB-12837/2020\|EPI_ISL_648483\|2020- | ···················· | ·A·················· | ····················· | ···················· | ···················· | ···················· |
| R | 1786 hCoV-19/USA/CA-CZB-12838/2020\|EPI_ISL_648484\|2020- | ···················· | ·A·················· | ····················· | ···················· | ···················· | ···················· |
| R | 1787 hCoV-19/USA/CA-CZB-12839/2020\|EPI_ISL_648485\|2020- | ···················· | ·A·················· | ····················· | ···················· | ···················· | ···················· |
| R | 1788 hCoV-19/USA/CA-CZB-12842/2020\|EPI_ISL_648487\|2020- | ···················· | ·A·················· | ····················· | ···················· | ···················· | ···················· |
| R | 1789 hCoV-19/USA/CA-CZB-12843/2020\|EPI_ISL_648488\|2020- | ···················· | ·A·················· | ····················· | ···················· | ···················· | ···················· |
| R | 1790 hCoV-19/USA/CA-CZB-12844/2020\|EPI_ISL_648489\|2020- | ···················· | ·A·················· | ····················· | ···················· | ···················· | ···················· |
| R | 1791 hCoV-19/USA/CA-CZB-7137/2020\|EPI_ISL_648497\|2020-0 | ···················· | ·A·················· | ····················· | ···················· | ···················· | ···················· |
| R | 1792 hCoV-19/USA/CA-CZB-7141/2020\|EPI_ISL_648498\|2020-0 | ···················· | ·A·················· | ····················· | ···················· | ···················· | ···················· |
| R | 1793 hCoV-19/England/QEUH-B0F7A7/2020\|EPI_ISL_641700\|20 | ···················· | ···················· | ····················· | ···················· | ···················· | ···················· |
| R | 1794 hCoV-19/England/QEUH-B0F09D/2020\|EPI_ISL_641701\|20 | ···················· | ···················· | ····················· | ···················· | ···················· | ···················· |
| R | 1795 hCoV-19/England/QEUH-B13C29/2020\|EPI_ISL_641702\|20 | ···················· | ···················· | ····················· | ···················· | ···················· | ···················· |
| R | 1796 hCoV-19/England/QEUH-B13C83/2020\|EPI_ISL_641703\|20 | ···················· | ···················· | ····················· | ···················· | ···················· | ···················· |
| R | 1797 hCoV-19/England/QEUH-B13EF6/2020\|EPI_ISL_641704\|20 | ···················· | ···················· | ····················· | ···················· | ···················· | ···················· |
| R | 1798 hCoV-19/England/QEUH-B14244/2020\|EPI_ISL_641705\|20 | ···················· | ···················· | ····················· | ···················· | ···················· | ···················· |
| R | 1799 hCoV-19/England/QEUH-B12CC0/2020\|EPI_ISL_641707\|20 | ···················· | ···················· | ····················· | ···················· | ···················· | ···················· |
| R | 1800 hCoV-19/England/QEUH-B14226/2020\|EPI_ISL_641708\|20 | ···················· | ···················· | ····················· | ···················· | ···················· | ···················· |
| R | 1801 hCoV-19/England/QEUH-B13E50/2020\|EPI_ISL_641709\|20 | ···················· | ···················· | ····················· | ···················· | ···················· | ···················· |
| R | 1802 hCoV-19/England/QEUH-B0F3A3/2020\|EPI_ISL_642200\|20 | ···················· | ···················· | ····················· | ···················· | ···················· | ···················· |
| R | 1803 hCoV-19/England/QEUH-B0F9DE/2020\|EPI_ISL_642201\|20 | ···················· | ···················· | ····················· | ···················· | ···················· | ···················· |
| R | 1804 hCoV-19/England/QEUH-B0E6AB/2020\|EPI_ISL_642203\|20 | ···················· | ···················· | ····················· | ···················· | ···················· | ···················· |
| R | 1805 hCoV-19/England/QEUH-B0DBAD/2020\|EPI_ISL_642205\|20 | ···················· | ···················· | ····················· | ···················· | ···················· | ···················· |
| R | 1806 hCoV-19/England/QEUH-B0E4A1/2020\|EPI_ISL_642206\|20 | ···················· | ···················· | ····················· | ···················· | ···················· | ···················· |
| R | 1807 hCoV-19/England/QEUH-B0DCB9/2020\|EPI_ISL_642207\|20 | ···················· | ···················· | ····················· | ···················· | ···················· | ···················· |
| R | 1808 hCoV-19/England/QEUH-B10521/2020\|EPI_ISL_642208\|20 | ···················· | ···················· | ····················· | ···················· | ···················· | ···················· |
| R | 1809 hCoV-19/England/QEUH-B11C1C/2020\|EPI_ISL_642209\|20 | ···················· | ·A·················· | ····················· | ···················· | ···················· | ··A················· |
| R | 1810 hCoV-19/England/QEUH-B12905/2020\|EPI_ISL_642211\|20 | ···················· | ···················· | ····················· | ···················· | ···················· | ···················· |
| R | 1811 hCoV-19/England/QEUH-B11818/2020\|EPI_ISL_642212\|20 | ···················· | ···················· | ····················· | ···················· | ···················· | ···················· |
| R | 1812 hCoV-19/England/QEUH-B10E17/2020\|EPI_ISL_642215\|20 | ···················· | ···················· | ····················· | ···················· | ···················· | ···················· |
| R | 1813 hCoV-19/England/QEUH-B10D83/2020\|EPI_ISL_642216\|20 | ···················· | ···················· | ····················· | ···················· | ···················· | ···················· |
| R | 1814 hCoV-19/England/QEUH-B11432/2020\|EPI_ISL_642218\|20 | ···················· | ···················· | ····················· | ···················· | ···················· | ···················· |
| R | 1815 hCoV-19/England/QEUH-B1197F/2020\|EPI_ISL_642220\|20 | ···················· | ···················· | ····················· | ···················· | ···················· | ···················· |
| R | 1816 hCoV-19/England/QEUH-B127A1/2020\|EPI_ISL_642221\|20 | ···················· | ···················· | ····················· | ···················· | ···················· | ···················· |
| R | 1817 hCoV-19/England/QEUH-B13139/2020\|EPI_ISL_642222\|20 | ···················· | ···················· | ····················· | ···················· | ···················· | ···················· |
| R | 1818 hCoV-19/England/QEUH-B11881/2020\|EPI_ISL_642223\|20 | ···················· | ···················· | ····················· | ···················· | ···················· | ···················· |
| R | 1819 hCoV-19/England/QEUH-B1298D/2020\|EPI_ISL_642225\|20 | ···················· | ···················· | ····················· | ···················· | ···················· | ···················· |
| R | 1820 hCoV-19/England/QEUH-B12194/2020\|EPI_ISL_642226\|20 | ···················· | ···················· | ····················· | ···················· | ···················· | ···················· |
| R | 1821 hCoV-19/England/QEUH-B11B88/2020\|EPI_ISL_642227\|20 | ···················· | ···················· | ····················· | ···················· | ···················· | ···················· |
| R | 1822 hCoV-19/England/QEUH-B10D1A/2020\|EPI_ISL_642228\|20 | ···················· | ···················· | ····················· | ···················· | ···················· | ···················· |
| R | 1823 hCoV-19/England/QEUH-B1206A/2020\|EPI_ISL_642229\|20 | ···················· | ···················· | ····················· | ···················· | ···················· | ···················· |
| R | 1824 hCoV-19/England/QEUH-B118EB/2020\|EPI_ISL_642230\|20 | ···················· | ···················· | ····················· | ···················· | ···················· | ···················· |
| R | 1825 hCoV-19/England/QEUH-B10F05/2020\|EPI_ISL_642231\|20 | ···················· | ···················· | ····················· | ···················· | ···················· | ···················· |
| R | 1826 hCoV-19/England/QEUH-B11C76/2020\|EPI_ISL_642232\|20 | ···················· | ···················· | ····················· | ···················· | ···················· | ···················· |
| R | 1827 hCoV-19/England/QEUH-B10F14/2020\|EPI_ISL_642233\|20 | ···················· | ···················· | ····················· | ···················· | ···················· | ···················· |
| R | 1828 hCoV-19/England/QEUH-B11511/2020\|EPI_ISL_642234\|20 | ···················· | ·A·················· | ····················· | ···················· | ···················· | ···················· |
| R | 1829 hCoV-19/England/QEUH-B10D65/2020\|EPI_ISL_642235\|20 | ···················· | ···················· | ····················· | ···················· | ···················· | ···················· |
| R | 1830 hCoV-19/England/QEUH-B1198E/2020\|EPI_ISL_642237\|20 | ···················· | ···················· | ····················· | ···················· | ···················· | ···················· |
| R | 1831 hCoV-19/England/QEUH-B11845/2020\|EPI_ISL_642238\|20 | ···················· | ···················· | ····················· | ···················· | ···················· | ···················· |
| R | 1832 hCoV-19/England/QEUH-B10C95/2020\|EPI_ISL_642240\|20 | ···················· | ···················· | ····················· | ···················· | ···················· | ···················· |
| R | 1833 hCoV-19/England/QEUH-B117C0/2020\|EPI_ISL_642241\|20 | ···················· | ···················· | ····················· | ···················· | ···················· | ···················· |
| R | 1834 hCoV-19/England/QEUH-B10EBD/2020\|EPI_ISL_642242\|20 | ···················· | ···················· | ····················· | ···················· | ···················· | ···················· |
| R | 1835 hCoV-19/England/QEUH-B10CE0/2020\|EPI_ISL_642244\|20 | ···················· | ···················· | ····················· | ···················· | ···················· | ···················· |
| R | 1836 hCoV-19/England/QEUH-B12923/2020\|EPI_ISL_642245\|20 | ···················· | ···················· | ····················· | ···················· | ···················· | ···················· |
| R | 1837 hCoV-19/England/QEUH-B13236/2020\|EPI_ISL_642246\|20 | ···················· | ···················· | ····················· | ···················· | ···················· | ···················· |
| R | 1838 hCoV-19/England/QEUH-B11B6A/2020\|EPI_ISL_642248\|20 | ···················· | ···················· | ····················· | ···················· | ···················· | ···················· |
| R | 1839 hCoV-19/England/QEUH-B115D5/2020\|EPI_ISL_642249\|20 | ···················· | ···················· | ····················· | ···················· | ···················· | ···················· |
| R | 1840 hCoV-19/England/QEUH-B1148D/2020\|EPI_ISL_642250\|20 | ···················· | ···················· | ····················· | ···················· | ···················· | ···················· |
| R | 1841 hCoV-19/England/QEUH-B12228/2020\|EPI_ISL_642251\|20 | ···················· | ···················· | ····················· | ···················· | ···················· | ···················· |
| R | 1842 hCoV-19/England/QEUH-B10DFC/2020\|EPI_ISL_642252\|20 | ···················· | ·A·················· | ····················· | ···················· | ···················· | ···················· |
| R | 1843 hCoV-19/England/QEUH-B11BF1/2020\|EPI_ISL_642253\|20 | ···················· | ···················· | ····················· | ···················· | ···················· | ···················· |
| R | 1844 hCoV-19/England/QEUH-B127FC/2020\|EPI_ISL_642254\|20 | ···················· | ···················· | ····················· | ···················· | ···················· | ···················· |
| R | 1845 hCoV-19/England/QEUH-B11BD3/2020\|EPI_ISL_642255\|20 | ···················· | ···················· | ····················· | ···················· | ···················· | ···················· |
| R | 1846 hCoV-19/England/QEUH-B11D55/2020\|EPI_ISL_642256\|20 | ···················· | ···················· | ····················· | ···················· | ···················· | ···················· |
| R | 1847 hCoV-19/England/QEUH-B10CC2/2020\|EPI_ISL_642257\|20 | ···················· | ···················· | ····················· | ···················· | ···················· | ···················· |
| R | 1848 hCoV-19/England/QEUH-B130D2/2020\|EPI_ISL_642259\|20 | ···················· | ···················· | ····················· | ···················· | ···················· | ···················· |
| R | 1849 hCoV-19/England/QEUH-B11BE2/2020\|EPI_ISL_642260\|20 | ···················· | ···················· | ····················· | ···················· | ···················· | ···················· |
| R | 1850 hCoV-19/England/QEUH-B11890/2020\|EPI_ISL_642261\|20 | ···················· | ···················· | ····················· | ···················· | ···················· | ···················· |
| R | 1851 hCoV-19/England/QEUH-B10E53/2020\|EPI_ISL_642262\|20 | ···················· | ···················· | ····················· | ···················· | ···················· | ···················· |
| R | 1852 hCoV-19/England/QEUH-B11A4F/2020\|EPI_ISL_642263\|20 | ···················· | ···················· | ····················· | ···················· | ···················· | ···················· |
| R | 1853 hCoV-19/England/QEUH-B129AB/2020\|EPI_ISL_642264\|20 | ···················· | ···················· | ····················· | ···················· | ···················· | ···················· |
| R | 1854 hCoV-19/England/QEUH-B11951/2020\|EPI_ISL_642265\|20 | ···················· | ···················· | ····················· | ···················· | ···················· | ···················· |
| R | 1855 hCoV-19/England/QEUH-B1146F/2020\|EPI_ISL_642266\|20 | ···················· | ·A·················· | ····················· | ···················· | ···················· | ···················· |

R 1856 hCoV-19/England/QEUH-B11687/2020|EPI_ISL_642268|20 ···················· ···················· ····················· ···················· ···················· ····················
R 1857 hCoV-19/England/QEUH-B12282/2020|EPI_ISL_642269|20 ···················· ···················· ····················· ···················· ···················· ····················
R 1858 hCoV-19/England/QEUH-B128AE/2020|EPI_ISL_642270|20 ···················· ···················· ····················· ···················· ···················· ····················
R 1859 hCoV-19/England/QEUH-B136A3/2020|EPI_ISL_642272|20 ···················· ···················· ····················· ···················· ···················· ····················
R 1860 hCoV-19/England/QEUH-B11C0D/2020|EPI_ISL_642273|20 ···················· ···················· ····················· ···················· ···················· ····················
R 1861 hCoV-19/England/QEUH-B10C77/2020|EPI_ISL_642275|20 ···················· ···················· ····················· ···················· ···················· ····················
R 1862 hCoV-19/England/QEUH-B11AE5/2020|EPI_ISL_642276|20 ···················· ···················· ····················· ···················· ···················· ····················
R 1863 hCoV-19/England/QEUH-B13148/2020|EPI_ISL_642277|20 ···················· ···················· ····················· ···················· ···················· ····················
R 1864 hCoV-19/England/QEUH-B13597/2020|EPI_ISL_642278|20 ···················· ···················· ····················· ···················· ···················· ····················
R 1865 hCoV-19/England/QEUH-B10F6F/2020|EPI_ISL_642279|20 ···················· ···················· ····················· ···················· ···················· ····················
R 1866 hCoV-19/England/QEUH-B13096/2020|EPI_ISL_642280|20 ···················· ·A·················· ····················· ···················· ···················· ···A················
R 1867 hCoV-19/England/QEUH-B126E0/2020|EPI_ISL_642281|20 ···················· ···················· ····················· ···················· ···················· ····················
R 1868 hCoV-19/England/QEUH-B1162D/2020|EPI_ISL_642282|20 ···················· ···················· ····················· ···················· ···················· ····················
R 1869 hCoV-19/England/QEUH-B1149C/2020|EPI_ISL_642283|20 ···················· ···················· ····················· ···················· ···················· ····················
R 1870 hCoV-19/England/QEUH-B13685/2020|EPI_ISL_642284|20 ···················· ···················· ····················· ···················· ···················· ····················
R 1871 hCoV-19/England/QEUH-B117EE/2020|EPI_ISL_642285|20 ···················· ···················· ····················· ···················· ···················· ····················
R 1872 hCoV-19/England/QEUH-B10EEA/2020|EPI_ISL_642286|20 ···················· ···················· ····················· ···················· ···················· ····················
R 1873 hCoV-19/England/QEUH-B12361/2020|EPI_ISL_642288|20 ···················· ···················· ····················· ···················· ···················· ····················
R 1874 hCoV-19/England/QEUH-B12835/2020|EPI_ISL_642290|20 ···················· ···················· ····················· ···················· ···················· ····················
R 1875 hCoV-19/England/QEUH-B104F7/2020|EPI_ISL_642291|20 ···················· ···················· ····················· ···················· ···················· ····················
R 1876 hCoV-19/England/QEUH-B11D64/2020|EPI_ISL_642292|20 ···················· ···················· ····················· ···················· ···················· ····················
R 1877 hCoV-19/England/QEUH-B11960/2020|EPI_ISL_642293|20 ···················· ···················· ····················· ···················· ···················· ····················
R 1878 hCoV-19/England/QEUH-B11BB5/2020|EPI_ISL_642294|20 ···················· ···················· ····················· ···················· ···················· ····················
R 1879 hCoV-19/England/QEUH-B119AC/2020|EPI_ISL_642295|20 ···················· ···················· ····················· ···················· ···················· ····················
R 1880 hCoV-19/England/QEUH-B11A5E/2020|EPI_ISL_642296|20 ···················· ···················· ····················· ···················· ···················· ····················
R 1881 hCoV-19/England/QEUH-B118FA/2020|EPI_ISL_642298|20 ···················· ···················· ····················· ···················· ···················· ····················
R 1882 hCoV-19/England/QEUH-B1271A/2020|EPI_ISL_642299|20 ···················· ···················· ····················· ···················· ···················· ····················
R 1883 hCoV-19/England/CAMC-B0C4D0/2020|EPI_ISL_643502|20 ···················· ···················· ····················· ···················· ···················· ····················
R 1884 hCoV-19/England/CAMC-B08B48/2020|EPI_ISL_643503|20 ···················· ···················· ····················· ···················· ···················· ····················
R 1885 hCoV-19/England/CAMC-B07E40/2020|EPI_ISL_643504|20 ···················· ···················· ····················· ···················· ···················· ····················
R 1886 hCoV-19/England/CAMC-B0B1DA/2020|EPI_ISL_643506|20 ···················· ···················· ····················· ···················· ···················· ····················
R 1887 hCoV-19/England/CAMC-B0AF1D/2020|EPI_ISL_643507|20 ···················· ·A·················· ····················· ···················· ···················· ···A················
R 1888 hCoV-19/England/CAMC-B08568/2020|EPI_ISL_643508|20 ···················· ···················· ····················· ···················· ···················· ····················
R 1889 hCoV-19/England/CAMC-B0898A/2020|EPI_ISL_643509|20 ···················· ···················· ····················· ···················· ···················· ····················
R 1890 hCoV-19/England/CAMC-B0B1F8/2020|EPI_ISL_643510|20 ···················· ···················· ····················· ···················· ···················· ····················
R 1891 hCoV-19/England/CAMC-B0CC50/2020|EPI_ISL_643511|20 ···················· ···················· ····················· ···················· ···················· ····················
R 1892 hCoV-19/England/CAMC-B0A8F4/2020|EPI_ISL_643512|20 ···················· ···················· ····················· ···················· ···················· ····················
R 1893 hCoV-19/England/CAMC-B0AAA3/2020|EPI_ISL_643514|20 ···················· ···················· ····················· ···················· ···················· ····················
R 1894 hCoV-19/England/CAMC-B0852C/2020|EPI_ISL_643515|20 ···················· ·A·················· ····················· ···················· ···················· ···A················
R 1895 hCoV-19/England/CAMC-B0AA67/2020|EPI_ISL_643516|20 ···················· ···················· ····················· ···················· ······T············· ····················
R 1896 hCoV-19/England/CAMC-B0B028/2020|EPI_ISL_643517|20 ···················· ·A·················· ····················· ···················· ···················· ···A················
R 1897 hCoV-19/England/CAMC-B0AD9B/2020|EPI_ISL_643518|20 ···················· ·A·················· ····················· ···················· ···················· ···A················
R 1898 hCoV-19/England/CAMC-B0C6AD/2020|EPI_ISL_643520|20 ···················· ···················· ····················· ···················· ···················· ····················
R 1899 hCoV-19/England/CAMC-B0AE2F/2020|EPI_ISL_643521|20 ···················· ···················· ····················· ···················· ···················· ····················
R 1900 hCoV-19/England/CAMC-B0CBF9/2020|EPI_ISL_643522|20 ···················· ···················· ····················· ···················· ···················· ····················
R 1901 hCoV-19/England/CAMC-B0801C/2020|EPI_ISL_643523|20 ···················· ···················· ····················· ···················· ···················· ····················
R 1902 hCoV-19/England/CAMC-B0C625/2020|EPI_ISL_643525|20 ···················· ···················· ····················· ···················· ···················· ····················
R 1903 hCoV-19/England/CAMC-B07DCB/2020|EPI_ISL_643526|20 ···················· ···················· ····················· ···················· ···················· ····················
R 1904 hCoV-19/England/CAMC-B0C4A3/2020|EPI_ISL_643527|20 ···················· ···················· ····················· ···················· ···················· ····················
R 1905 hCoV-19/England/CAMC-B07E04/2020|EPI_ISL_643528|20 ···················· ···················· ····················· ···················· ···················· ····················
R 1906 hCoV-19/England/CAMC-B08216/2020|EPI_ISL_643529|20 ···················· ···················· ····················· ···················· ······T············· ···············A····
R 1907 hCoV-19/England/CAMC-B084F2/2020|EPI_ISL_643530|20 ···················· ···················· ····················· ···················· ···················· ····················
R 1908 hCoV-19/England/CAMC-B0B42C/2020|EPI_ISL_643531|20 ···················· ·A·················· ····················· ···················· ···················· ····················
R 1909 hCoV-19/England/CAMC-B0AE7A/2020|EPI_ISL_643532|20 ···················· ···················· ····················· ···················· ···················· ····················
R 1910 hCoV-19/England/CAMC-B0B705/2020|EPI_ISL_643533|20 ···················· ···················· ····················· ···················· ···················· ····················
R 1911 hCoV-19/England/CAMC-B07AA6/2020|EPI_ISL_643534|20 ···················· ···················· ····················· ···················· ···················· ····················
R 1912 hCoV-19/England/CAMC-B0853B/2020|EPI_ISL_643535|20 ···················· ···················· ····················· ···················· ···················· ····················
R 1913 hCoV-19/England/CAMC-B0AE98/2020|EPI_ISL_643536|20 ···················· ·A·················· ····················· ···················· ···················· ···A················
R 1914 hCoV-19/England/CAMC-B084B6/2020|EPI_ISL_643537|20 ···················· ···················· ····················· ···················· ···················· ····················
R 1915 hCoV-19/England/CAMC-B08225/2020|EPI_ISL_643538|20 ···················· ···················· ····················· ···················· ···················· ····················
R 1916 hCoV-19/England/CAMC-B07E6E/2020|EPI_ISL_643539|20 ···················· ···················· ····················· ···················· ···················· ····················
R 1917 hCoV-19/England/CAMC-B0D09F/2020|EPI_ISL_643540|20 ···················· ···················· ····················· ···················· ···················· ····················
R 1918 hCoV-19/England/CAMC-B0CD99/2020|EPI_ISL_643541|20 ···················· ·A·················· ····················· ···················· ···················· ···A················
R 1919 hCoV-19/England/CAMC-B0B78D/2020|EPI_ISL_643542|20 ···················· ···················· ····················· ···················· ···················· ····················
R 1920 hCoV-19/England/CAMC-B0B4E0/2020|EPI_ISL_643543|20 ···················· ···················· ····················· ···················· ···················· ····················
R 1921 hCoV-19/England/CAMC-B0D071/2020|EPI_ISL_643544|20 ···················· ···················· ····················· ···················· ···················· ····················
R 1922 hCoV-19/England/CAMC-B08094/2020|EPI_ISL_643545|20 ···················· ···················· ····················· ···················· ······T············· ····················
R 1923 hCoV-19/England/CAMC-B07AB5/2020|EPI_ISL_643546|20 ···················· ·A·················· ····················· ···················· ···················· ···A················
R 1924 hCoV-19/England/CAMC-B0CBCC/2020|EPI_ISL_643547|20 ···················· ···················· ····················· ···················· ···················· ····················
R 1925 hCoV-19/England/CAMC-B0B0A0/2020|EPI_ISL_643548|20 ···················· ·A·················· ····················· ···················· ···················· ···A················
R 1926 hCoV-19/England/CAMC-B089B7/2020|EPI_ISL_643549|20 ···················· ···················· ····················· ···················· ···················· ····················
R 1927 hCoV-19/England/CAMC-B0C92C/2020|EPI_ISL_643550|20 ···················· ···················· ····················· ···················· ···················· ····················
R 1928 hCoV-19/England/CAMC-B0B477/2020|EPI_ISL_643551|20 ···················· ···················· ····················· ···················· ···················· ····················
R 1929 hCoV-19/England/CAMC-B0B398/2020|EPI_ISL_643552|20 ···················· ···················· ····················· ···················· ···················· ····················
R 1930 hCoV-19/England/CAMC-B0B714/2020|EPI_ISL_643553|20 ···················· ···················· ····················· ···················· ···················· ····················
R 1931 hCoV-19/England/CAMC-B07E22/2020|EPI_ISL_643554|20 ···················· ···················· ····················· ···················· ···················· ····················
R 1932 hCoV-19/England/CAMC-B0B3D4/2020|EPI_ISL_643555|20 ···················· ···················· ····················· ···················· ···················· ····················
R 1933 hCoV-19/England/CAMC-B0CD4E/2020|EPI_ISL_643556|20 ···················· ···················· ····················· ···················· ···················· ····················
R 1934 hCoV-19/England/CAMC-B0B301/2020|EPI_ISL_643557|20 ···················· ···················· ····················· ···················· ······T············· ····················
R 1935 hCoV-19/England/CAMC-B0C8E3/2020|EPI_ISL_643558|20 ···················· ···················· ····················· ···················· ···················· ····················

| | | | | | | | |
|---|---|---|---|---|---|---|---|
| R | 1936 hCoV-19/England/CAMC-B0850E/2020\|EPI_ISL_643559\|20 | .................... | .................... | .................... | .................... | .................... | .................... |
| R | 1937 hCoV-19/England/CAMC-B0B5ED/2020\|EPI_ISL_643560\|20 | .................... | .................... | .................... | .................... | .................... | .................... |
| R | 1938 hCoV-19/England/CAMC-B083B9/2020\|EPI_ISL_643561\|20 | .................... | .................... | .................... | .................... | .................... | .................... |
| R | 1939 hCoV-19/England/CAMC-B0B116/2020\|EPI_ISL_643562\|20 | .................... | .................... | .................... | .................... | .................... | .................... |
| R | 1940 hCoV-19/England/CAMC-B08489/2020\|EPI_ISL_643563\|20 | .................... | .................... | .................... | .................... | .................... | .................... |
| R | 1941 hCoV-19/England/CAMC-B0AC43/2020\|EPI_ISL_643564\|20 | .................... | .................... | .................... | .................... | .................... | .................... |
| R | 1942 hCoV-19/England/CAMC-B07A97/2020\|EPI_ISL_643565\|20 | .................... | .................... | .................... | .................... | .................... | .................... |
| R | 1943 hCoV-19/England/CAMC-B0B3F2/2020\|EPI_ISL_643566\|20 | .................... | .................... | .................... | .................... | .................... | .................... |
| R | 1944 hCoV-19/England/CAMC-B0B125/2020\|EPI_ISL_643567\|20 | .................... | .A.................. | .................... | .................... | .................... | ..A................. |
| R | 1945 hCoV-19/England/CAMC-B07A88/2020\|EPI_ISL_643568\|20 | .................... | .................... | .................... | .................... | .................... | .................... |
| R | 1946 hCoV-19/England/CAMC-B0C634/2020\|EPI_ISL_643569\|20 | .................... | .................... | .................... | .................... | .................... | .................... |
| R | 1947 hCoV-19/England/CAMC-B0CD5D/2020\|EPI_ISL_643571\|20 | .................... | .................... | .................... | .................... | .................... | .................... |
| R | 1948 hCoV-19/England/CAMC-B0AF3B/2020\|EPI_ISL_643573\|20 | .................... | .................... | .................... | .................... | .................... | .................... |
| R | 1949 hCoV-19/England/CAMC-B0AB0A/2020\|EPI_ISL_643574\|20 | .................... | .................... | .................... | .................... | .................... | .................... |
| R | 1950 hCoV-19/England/CAMC-B0BAA2/2020\|EPI_ISL_643575\|20 | .................... | .................... | .................... | .................... | .................... | .................... |
| R | 1951 hCoV-19/England/CAMC-B0AFFF/2020\|EPI_ISL_643576\|20 | .................... | .................... | .................... | .................... | .................... | .................... |
| R | 1952 hCoV-19/England/CAMC-B0B1AD/2020\|EPI_ISL_643577\|20 | .................... | .................... | .................... | .................... | ......T............. | .................... |
| R | 1953 hCoV-19/England/CAMC-B084E3/2020\|EPI_ISL_643579\|20 | .................... | .................... | .................... | .................... | .................... | .................... |
| R | 1954 hCoV-19/England/CAMC-B08252/2020\|EPI_ISL_643580\|20 | .................... | .................... | .................... | .................... | .................... | .................... |
| R | 1955 hCoV-19/England/CAMC-B07EAA/2020\|EPI_ISL_643581\|20 | .................... | .................... | .................... | .................... | .................... | .................... |
| R | 1956 hCoV-19/England/CAMC-B0C82F/2020\|EPI_ISL_643582\|20 | .................... | .................... | .................... | .................... | .................... | .................... |
| R | 1957 hCoV-19/England/CAMC-B0C83E/2020\|EPI_ISL_643583\|20 | .................... | .................... | .................... | .................... | .................... | .................... |
| R | 1958 hCoV-19/England/CAMC-B08586/2020\|EPI_ISL_643584\|20 | .................... | .................... | .................... | .................... | .................... | .................... |
| R | 1959 hCoV-19/England/CAMC-B0C652/2020\|EPI_ISL_643585\|20 | .................... | .................... | .................... | .................... | .................... | .................... |
| R | 1960 hCoV-19/England/CAMC-B08067/2020\|EPI_ISL_643586\|20 | .................... | .................... | .................... | .................... | .................... | .................... |
| R | 1961 hCoV-19/England/CAMC-B0845C/2020\|EPI_ISL_643587\|20 | .................... | .................... | .................... | .................... | .................... | .................... |
| R | 1962 hCoV-19/England/CAMC-B0B4A4/2020\|EPI_ISL_643588\|20 | .................... | .................... | .................... | .................... | .................... | .................... |
| R | 1963 hCoV-19/England/CAMC-B0C87A/2020\|EPI_ISL_643589\|20 | .................... | .................... | .................... | .................... | .................... | .................... |
| R | 1964 hCoV-19/England/CAMC-B0C42B/2020\|EPI_ISL_643590\|20 | .................... | .................... | .................... | .................... | .................... | .................... |
| R | 1965 hCoV-19/England/CAMC-B0AAFE/2020\|EPI_ISL_643591\|20 | .................... | .................... | .................... | .................... | .................... | .................... |
| R | 1966 hCoV-19/England/CAMC-B0CD8A/2020\|EPI_ISL_643592\|20 | .................... | .................... | .................... | .................... | .................... | .................... |
| R | 1967 hCoV-19/England/CAMC-B08049/2020\|EPI_ISL_643593\|20 | .................... | .................... | .................... | .................... | .................... | .................... |
| R | 1968 hCoV-19/England/CAMC-B0897B/2020\|EPI_ISL_643594\|20 | .................... | .................... | .................... | .................... | .................... | .................... |
| R | 1969 hCoV-19/England/CAMC-B0B2E6/2020\|EPI_ISL_643595\|20 | .................... | .................... | .................... | .................... | .................... | .................... |
| R | 1970 hCoV-19/England/CAMC-B0C616/2020\|EPI_ISL_643596\|20 | .................... | .................... | .................... | .................... | .................... | .................... |
| R | 1971 hCoV-19/England/CAMC-B0CCBA/2020\|EPI_ISL_643597\|20 | .................... | .................... | .................... | .................... | .................... | .................... |
| R | 1972 hCoV-19/England/CAMC-B07B49/2020\|EPI_ISL_643598\|20 | .................... | .................... | .................... | .................... | .................... | .................... |
| R | 1973 hCoV-19/England/CAMC-B0C93B/2020\|EPI_ISL_643599\|20 | .................... | .................... | .................... | .................... | .................... | .................... |
| R | 1974 hCoV-19/USA/VA-DCLS-1662/2020\|EPI_ISL_644833\|2020- | .................... | .A.................. | .................... | .................... | .................... | .................... |
| R | 1975 hCoV-19/USA/VA-DCLS-1663/2020\|EPI_ISL_644834\|2020- | .................... | .................... | .................... | .................... | .................... | .................... |
| R | 1976 hCoV-19/USA/VA-DCLS-1664/2020\|EPI_ISL_644835\|2020- | .................... | .................... | .................... | .................... | .................... | .................... |
| R | 1977 hCoV-19/USA/VA-DCLS-1665/2020\|EPI_ISL_644836\|2020- | .................... | .................... | .................... | .................... | .................... | .................... |
| R | 1978 hCoV-19/USA/VA-DCLS-1666/2020\|EPI_ISL_644837\|2020- | .................... | .................... | .................... | .................... | .................... | .................... |
| R | 1979 hCoV-19/USA/VA-DCLS-1671/2020\|EPI_ISL_644839\|2020- | .................... | .A.................. | .................... | .................... | .................... | .................... |
| R | 1980 hCoV-19/USA/VA-DCLS-1672/2020\|EPI_ISL_644840\|2020- | .................... | .................... | .................... | .................... | .................... | .................... |
| R | 1981 hCoV-19/USA/VA-DCLS-1673/2020\|EPI_ISL_644841\|2020- | .................... | .................... | .................... | .................... | .................... | .................... |
| R | 1982 hCoV-19/USA/VA-DCLS-1736/2020\|EPI_ISL_644842\|2020- | .................... | .................... | .................... | .................... | .................... | .................... |
| R | 1983 hCoV-19/USA/VA-DCLS-1737/2020\|EPI_ISL_644843\|2020- | .................... | .................... | .................... | .................... | .................... | .................... |
| R | 1984 hCoV-19/USA/VA-DCLS-1739/2020\|EPI_ISL_644844\|2020- | .................... | .................... | .................... | .................... | .................... | .................... |
| R | 1985 hCoV-19/USA/VA-DCLS-1740/2020\|EPI_ISL_644845\|2020- | .................... | .................... | .................... | .................... | .................... | .................... |
| R | 1986 hCoV-19/USA/VA-DCLS-1743/2020\|EPI_ISL_644846\|2020- | .................... | .A.................. | .................... | .................... | .................... | .................... |
| R | 1987 hCoV-19/USA/VA-DCLS-1744/2020\|EPI_ISL_644847\|2020- | .................... | .A.................. | .................... | .................... | .................... | .................... |
| R | 1988 hCoV-19/USA/VA-DCLS-1745/2020\|EPI_ISL_644848\|2020- | .................... | .................... | .................... | .................... | .................... | .................... |
| R | 1989 hCoV-19/USA/VA-DCLS-1746/2020\|EPI_ISL_644849\|2020- | .................... | .................... | .................... | .................... | .................... | .................... |
| R | 1990 hCoV-19/USA/VA-DCLS-1749/2020\|EPI_ISL_644850\|2020- | .................... | .................... | .................... | .................... | .................... | .................... |
| R | 1991 hCoV-19/USA/VA-DCLS-1750/2020\|EPI_ISL_644851\|2020- | .................... | .................... | .................... | .................... | .................... | .................... |
| R | 1992 hCoV-19/USA/VA-DCLS-1752/2020\|EPI_ISL_644852\|2020- | .................... | .................... | .................... | .................... | .................... | .................... |
| R | 1993 hCoV-19/USA/VA-DCLS-1753/2020\|EPI_ISL_644853\|2020- | .................... | .A.................. | .................... | .................... | .................... | .................... |
| R | 1994 hCoV-19/USA/VA-DCLS-1755/2020\|EPI_ISL_644854\|2020- | .................... | ....A............... | .................... | .................... | .................... | .................... |
| R | 1995 hCoV-19/USA/VA-DCLS-1758/2020\|EPI_ISL_644855\|2020- | .................... | .................... | .................... | .................... | .................... | .................... |
| R | 1996 hCoV-19/USA/VA-DCLS-1761/2020\|EPI_ISL_644856\|2020- | .................... | .................... | .................... | .................... | .................... | .................... |
| R | 1997 hCoV-19/USA/VA-DCLS-1762/2020\|EPI_ISL_644857\|2020- | .................... | .A.................. | .................... | .................... | .................... | .................... |
| R | 1998 hCoV-19/USA/VA-DCLS-1764/2020\|EPI_ISL_644858\|2020- | .................... | .................... | .................... | .................... | .................... | .................... |
| R | 1999 hCoV-19/USA/VA-DCLS-1765/2020\|EPI_ISL_644859\|2020- | .................... | .................... | .................... | .................... | .................... | .................... |
| R | 2000 hCoV-19/USA/VA-DCLS-1766/2020\|EPI_ISL_644860\|2020- | .................... | .................... | .................... | .................... | .................... | .................... |
| R | 2001 hCoV-19/USA/VA-DCLS-1767/2020\|EPI_ISL_644861\|2020- | .................... | .................... | .................... | .................... | .................... | .................... |
| R | 2002 hCoV-19/USA/VA-DCLS-1769/2020\|EPI_ISL_644862\|2020- | .................... | .............A...... | .................... | .................... | .................... | .................... |
| R | 2003 hCoV-19/USA/VA-DCLS-1770/2020\|EPI_ISL_644863\|2020- | .................... | .................... | .................... | .................... | .................... | .................... |
| R | 2004 hCoV-19/USA/VA-DCLS-1772/2020\|EPI_ISL_644864\|2020- | .................... | .................... | .................... | .................... | .................... | .................... |
| R | 2005 hCoV-19/USA/VA-DCLS-1775/2020\|EPI_ISL_644865\|2020- | .................... | .................... | .................... | .................... | .................... | .................... |
| R | 2006 hCoV-19/USA/VA-DCLS-1776/2020\|EPI_ISL_644866\|2020- | .................... | .................... | .................... | .................... | .................... | .................... |
| R | 2007 hCoV-19/USA/VA-DCLS-1777/2020\|EPI_ISL_644867\|2020- | .................... | .................... | .................... | .................... | .................... | .................... |
| R | 2008 hCoV-19/USA/VA-DCLS-1778/2020\|EPI_ISL_644868\|2020- | .................... | .................... | .................... | .................... | .................... | .................... |
| R | 2009 hCoV-19/USA/VA-DCLS-1783/2020\|EPI_ISL_644869\|2020- | .................... | .................... | .................... | .................... | .................... | .................... |
| R | 2010 hCoV-19/USA/VA-DCLS-1784/2020\|EPI_ISL_644870\|2020- | .................... | .................... | .................... | .................... | .................... | .................... |
| R | 2011 hCoV-19/USA/VA-DCLS-1785/2020\|EPI_ISL_644871\|2020- | .................... | .................... | .................... | .................... | .................... | .................... |
| R | 2012 hCoV-19/USA/VA-DCLS-1786/2020\|EPI_ISL_644872\|2020- | .................... | .............A...... | .................... | .................... | .................... | .................... |
| R | 2013 hCoV-19/USA/VA-DCLS-1788/2020\|EPI_ISL_644873\|2020- | .................... | .A.................. | .................... | .................... | .................... | .................... |
| R | 2014 hCoV-19/USA/VA-DCLS-1790/2020\|EPI_ISL_644874\|2020- | .................... | .A.................. | .................... | .................... | .................... | .................... |
| R | 2015 hCoV-19/USA/VA-DCLS-1791/2020\|EPI_ISL_644875\|2020- | .................... | .................... | .................... | .................... | .................... | .................... |

| | | | | | | | |
|---|---|---|---|---|---|---|---|
| R | 2016 hCoV-19/USA/VA-DCLS-1793/2020\|EPI_ISL_644876\|2020- | ···················· | ·A·················· | ····················· | ···················· | ···················· | ···················· |
| R | 2017 hCoV-19/USA/VA-DCLS-1794/2020\|EPI_ISL_644877\|2020- | ···················· | ···················· | ····················· | ···················· | ···················· | ···················· |
| R | 2018 hCoV-19/USA/VA-DCLS-1795/2020\|EPI_ISL_644878\|2020- | ···················· | ···················· | ····················· | ···················· | ···················· | ···················· |
| R | 2019 hCoV-19/USA/VA-DCLS-1796/2020\|EPI_ISL_644879\|2020- | ···················· | ···················· | ····················· | ···················· | ···················· | ···················· |
| R | 2020 hCoV-19/USA/VA-DCLS-1798/2020\|EPI_ISL_644880\|2020- | ···················· | ···················· | ····················· | ···················· | ···················· | ···················· |
| R | 2021 hCoV-19/USA/VA-DCLS-1800/2020\|EPI_ISL_644881\|2020- | ···················· | ···················· | ····················· | ···················· | ···················· | ···················· |
| R | 2022 hCoV-19/USA/VA-DCLS-1956/2020\|EPI_ISL_644882\|2020- | ···················· | ···················· | ····················· | ···················· | ···················· | ···················· |
| R | 2023 hCoV-19/USA/VA-DCLS-1960/2020\|EPI_ISL_644883\|2020- | ···················· | ···················· | ····················· | ···················· | ···················· | ···················· |
| R | 2024 hCoV-19/USA/VA-DCLS-1961/2020\|EPI_ISL_644884\|2020- | ···················· | ···················· | ····················· | ···················· | ···················· | ···················· |
| R | 2025 hCoV-19/USA/VA-DCLS-1963/2020\|EPI_ISL_644885\|2020- | ···················· | ·A·················· | ····················· | ···················· | ···················· | ···················· |
| R | 2026 hCoV-19/USA/VA-DCLS-1967/2020\|EPI_ISL_644886\|2020- | ···················· | ·A·················· | ····················· | ···················· | ···················· | ···················· |
| R | 2027 hCoV-19/USA/VA-DCLS-1969/2020\|EPI_ISL_644887\|2020- | ···················· | ···················· | ····················· | ···················· | ···················· | ···················· |
| R | 2028 hCoV-19/USA/VA-DCLS-1971/2020\|EPI_ISL_644888\|2020- | ···················· | ·A·················· | ····················· | ···················· | ···················· | ···················· |
| R | 2029 hCoV-19/USA/VA-DCLS-1972/2020\|EPI_ISL_644889\|2020- | ···················· | ···················· | ····················· | ···················· | ···················· | ···················· |
| R | 2030 hCoV-19/USA/VA-DCLS-1975/2020\|EPI_ISL_644891\|2020- | ···················· | ···················· | ····················· | ···················· | ···················· | ···················· |
| R | 2031 hCoV-19/USA/VA-DCLS-1977/2020\|EPI_ISL_644892\|2020- | ···················· | ···················· | ····················· | ···················· | ···················· | ···················· |
| R | 2032 hCoV-19/USA/VA-DCLS-1979/2020\|EPI_ISL_644894\|2020- | ···················· | ···················· | ····················· | ···················· | ···················· | ···················· |
| R | 2033 hCoV-19/USA/VA-DCLS-1980/2020\|EPI_ISL_644895\|2020- | ···················· | ···················· | ····················· | ···················· | ···················· | ···················· |
| R | 2034 hCoV-19/USA/VA-DCLS-1981/2020\|EPI_ISL_644896\|2020- | ···················· | ···················· | ····················· | ···················· | ···················· | ···················· |
| R | 2035 hCoV-19/USA/VA-DCLS-1983/2020\|EPI_ISL_644898\|2020- | ···················· | ···················· | ····················· | ···················· | ···················· | ···················· |
| R | 2036 hCoV-19/USA/VA-DCLS-1984/2020\|EPI_ISL_644899\|2020- | ···················· | ···················· | ····················· | ···················· | ···················· | ···················· |
| R | 2037 hCoV-19/England/CAMC-B21DDA/2020\|EPI_ISL_647160\|20 | ···················· | ···················· | ····················· | ···················· | ···················· | ···················· |
| R | 2038 hCoV-19/England/CAMC-B21C64/2020\|EPI_ISL_647161\|20 | ···················· | ···················· | ····················· | ···················· | ······T············· | ···················· |
| R | 2039 hCoV-19/England/CAMC-B21332/2020\|EPI_ISL_647163\|20 | ···················· | ···················· | ····················· | ···················· | ···················· | ···················· |
| R | 2040 hCoV-19/England/CAMC-B22823/2020\|EPI_ISL_647165\|20 | ···················· | ·A·················· | ····················· | ···················· | ···················· | ···················· |
| R | 2041 hCoV-19/England/CAMC-B21754/2020\|EPI_ISL_647168\|20 | ···················· | ·A·················· | ····················· | ···················· | ···················· | ···A················ |
| R | 2042 hCoV-19/England/CAMC-B21A4C/2020\|EPI_ISL_647169\|20 | ···················· | ···················· | ····················· | ···················· | ···················· | ···················· |
| R | 2044 hCoV-19/USA/CA-CZB-12852/2020\|EPI_ISL_648493\|2020- | ···················· | ·A·················· | ····················· | ···················· | ···················· | ···················· |
| R | 2045 hCoV-19/USA/CA-CZB-12854/2020\|EPI_ISL_648494\|2020- | ···················· | ·A·················· | ····················· | ···················· | ···················· | ···················· |
| R | 2046 hCoV-19/USA/CA-CZB-12855/2020\|EPI_ISL_648495\|2020- | ···················· | ·A·················· | ····················· | ···················· | ···················· | ···················· |
| R | 2048 hCoV-19/USA/CA-CZB-10291/2020\|EPI_ISL_648499\|2020- | ···················· | ·A·················· | ····················· | ···················· | ···················· | ···················· |
| R | 2049 hCoV-19/Australia/VIC12506/2020\|EPI_ISL_640358\|202 | ···················· | ···················· | ····················· | ···················· | ···················· | ···················· |
| R | 2051 hCoV-19/Australia/VIC12879/2020\|EPI_ISL_640360\|202 | ···················· | ···················· | ····················· | ···················· | ···················· | ···················· |
| R | 2052 hCoV-19/Australia/VIC13253/2020\|EPI_ISL_640361\|202 | ···················· | ···················· | ····················· | ···················· | ···················· | ···················· |
| R | 2054 hCoV-19/Australia/VIC16714/2020\|EPI_ISL_640378\|202 | ···················· | ···················· | ····················· | ···················· | ···················· | ···················· |
| R | 2055 hCoV-19/Australia/VIC12265/2020\|EPI_ISL_640379\|202 | ···················· | ···················· | ····················· | ···················· | ···················· | ···················· |
| R | 2056 hCoV-19/Australia/VIC12145/2020\|EPI_ISL_640380\|202 | ···················· | ······A············· | ····················· | ···················· | ···················· | ···················· |
| R | 2057 hCoV-19/Australia/VIC15972/2020\|EPI_ISL_640382\|202 | ···················· | ···················· | ····················· | ···················· | ···················· | ···················· |
| R | 2058 hCoV-19/Australia/VIC11716/2020\|EPI_ISL_640383\|202 | ···················· | ···················· | ····················· | ···················· | ···················· | ···················· |
| R | 2059 hCoV-19/Australia/VIC12268/2020\|EPI_ISL_640384\|202 | ···················· | ···················· | ····················· | ···················· | ···················· | ···················· |
| R | 2061 hCoV-19/Australia/VIC12160/2020\|EPI_ISL_640388\|202 | ···················· | ···················· | ····················· | ···················· | ···················· | ···················· |
| R | 2062 hCoV-19/Australia/VIC16135/2020\|EPI_ISL_640390\|202 | ···················· | ···················· | ····················· | ···················· | ···················· | ···················· |
| R | 2063 hCoV-19/Australia/VIC11815/2020\|EPI_ISL_640391\|202 | ···················· | ···················· | ···K················· | ···················· | ···················· | ···················· |
| R | 2064 hCoV-19/Australia/VIC15914/2020\|EPI_ISL_640392\|202 | ···················· | ···················· | ····················· | ···················· | ···················· | ···················· |
| R | 2065 hCoV-19/Australia/VIC12538/2020\|EPI_ISL_640394\|202 | ···················· | ···················· | ····················· | ···················· | ···················· | ···················· |
| R | 2066 hCoV-19/Australia/VIC12607/2020\|EPI_ISL_640396\|202 | ···················· | ···················· | ····················· | ···················· | ···················· | ···················· |
| R | 2067 hCoV-19/Australia/VIC12471/2020\|EPI_ISL_640397\|202 | ···················· | ···················· | ····················· | ···················· | ···················· | ···················· |
| R | 2068 hCoV-19/Australia/VIC16569/2020\|EPI_ISL_640398\|202 | ···················· | ···················· | ····················· | ···················· | ···················· | ···················· |
| R | 2069 hCoV-19/Australia/VIC16654/2020\|EPI_ISL_640400\|202 | ···················· | ···················· | ····················· | ···················· | ···················· | ···················· |
| R | 2071 hCoV-19/Australia/VIC12266/2020\|EPI_ISL_640402\|202 | ···················· | ···················· | ····················· | ···················· | ···················· | ···················· |
| R | 2072 hCoV-19/Australia/VIC13272/2020\|EPI_ISL_640403\|202 | ···················· | ···················· | ····················· | ···················· | ···················· | ···················· |
| R | 2073 hCoV-19/Australia/VIC16136/2020\|EPI_ISL_640404\|202 | ···················· | ···················· | ····················· | ···················· | ···················· | ···················· |
| R | 2074 hCoV-19/Australia/VIC12674/2020\|EPI_ISL_640405\|202 | ···················· | ···················· | ····················· | ···················· | ···················· | ···················· |
| R | 2077 hCoV-19/Australia/VIC12665/2020\|EPI_ISL_640408\|202 | ···················· | ···················· | ····················· | ···················· | ···················· | ···················· |
| R | 2078 hCoV-19/Australia/VIC12142/2020\|EPI_ISL_640410\|202 | ···················· | ···················· | ····················· | ···················· | ···················· | ···················· |
| R | 2079 hCoV-19/Australia/VIC12198/2020\|EPI_ISL_640411\|202 | ···················· | ···················· | ····················· | ···················· | ···················· | ···················· |
| R | 2080 hCoV-19/Australia/VIC16573/2020\|EPI_ISL_640412\|202 | ···················· | ···················· | ····················· | ···················· | ···················· | ···················· |
| R | 2081 hCoV-19/Australia/VIC16347/2020\|EPI_ISL_640413\|202 | ···················· | ···················· | ····················· | ···················· | ···················· | ···················· |
| R | 2082 hCoV-19/Australia/VIC16680/2020\|EPI_ISL_640415\|202 | ···················· | ···················· | ····················· | ···················· | ···················· | ···················· |
| R | 2083 hCoV-19/Australia/VIC12767/2020\|EPI_ISL_640416\|202 | ···················· | ···················· | ····················· | ···················· | ···················· | ···················· |
| R | 2084 hCoV-19/Australia/VIC12375/2020\|EPI_ISL_640417\|202 | ···················· | ···················· | ····················· | ···················· | ···················· | ···················· |
| R | 2085 hCoV-19/Australia/VIC12433/2020\|EPI_ISL_640419\|202 | ···················· | ···················· | ····················· | ···················· | ···················· | ···················· |
| R | 2086 hCoV-19/Australia/VIC16350/2020\|EPI_ISL_640420\|202 | ···················· | ···················· | ····················· | ···················· | ···················· | ···················· |
| R | 2087 hCoV-19/Australia/VIC12686/2020\|EPI_ISL_640421\|202 | ···················· | ···················· | ····················· | ···················· | ···················· | ···················· |
| R | 2089 hCoV-19/Australia/VIC12393/2020\|EPI_ISL_640423\|202 | ···················· | ···················· | ····················· | ···················· | ···················· | ···················· |
| R | 2090 hCoV-19/Australia/VIC16039/2020\|EPI_ISL_640424\|202 | ···················· | ···················· | ····················· | ···················· | ···················· | ···················· |
| R | 2091 hCoV-19/Australia/VIC16380/2020\|EPI_ISL_640425\|202 | ···················· | ···················· | ····················· | ···················· | ···················· | ···················· |
| R | 2092 hCoV-19/Australia/VIC16153/2020\|EPI_ISL_640427\|202 | ···················· | ···················· | ····················· | ···················· | ···················· | ···················· |
| R | 2093 hCoV-19/Australia/VIC16407/2020\|EPI_ISL_640432\|202 | ···················· | ···················· | ····················· | ···················· | ···················· | ···················· |
| R | 2094 hCoV-19/Australia/VIC12412/2020\|EPI_ISL_640433\|202 | ···················· | ···················· | ····················· | ···················· | ···················· | ···················· |
| R | 2095 hCoV-19/Australia/VIC12444/2020\|EPI_ISL_640434\|202 | ···················· | ···················· | ····················· | ···················· | ···················· | ···················· |
| R | 2096 hCoV-19/Australia/VIC12151/2020\|EPI_ISL_640435\|202 | ···················· | ···················· | ····················· | ···················· | ···················· | ···················· |
| R | 2097 hCoV-19/Australia/VIC16488/2020\|EPI_ISL_640436\|202 | ···················· | ···················· | ····················· | ···················· | ···················· | ···················· |
| R | 2098 hCoV-19/Australia/VIC12121/2020\|EPI_ISL_640437\|202 | ···················· | ···················· | ····················· | ···················· | ···················· | ···················· |
| R | 2099 hCoV-19/Australia/VIC13260/2020\|EPI_ISL_640439\|202 | ···················· | ···················· | ····················· | ···················· | ···················· | ···················· |
| R | 2100 hCoV-19/Australia/VIC16095/2020\|EPI_ISL_640440\|202 | ···················· | ···················· | ····················· | ···················· | ···················· | ···················· |
| R | 2101 hCoV-19/Australia/VIC12553/2020\|EPI_ISL_640441\|202 | ···················· | ···················· | ····················· | ···················· | ···················· | ···················· |
| R | 2102 hCoV-19/Australia/VIC16106/2020\|EPI_ISL_640442\|202 | ···················· | ···················· | ····················· | ···················· | ···················· | ···················· |
| R | 2103 hCoV-19/Australia/VIC11974/2020\|EPI_ISL_640444\|202 | ···················· | ···················· | ····················· | ···················· | ···················· | ···················· |
| R | 2104 hCoV-19/Australia/VIC16565/2020\|EPI_ISL_640445\|202 | ···················· | ···················· | ····················· | ···················· | ···················· | ···················· |

| | | | | | | | |
|---|---|---|---|---|---|---|---|
| R | 2105 hCoV-19/Australia/VIC12690/2020\|EPI_ISL_640446\|202 | ···················· | ···················· | ····················· | ···················· | ···················· | ···················· |
| R | 2107 hCoV-19/Australia/VIC11958/2020\|EPI_ISL_640449\|202 | ···················· | ···················· | ····················· | ···················· | ···················· | ···················· |
| R | 2108 hCoV-19/Australia/VIC12726/2020\|EPI_ISL_640450\|202 | ···················· | ···················· | ····················· | ···················· | ···················· | ···················· |
| R | 2109 hCoV-19/Australia/VIC12047/2020\|EPI_ISL_640451\|202 | ···················· | ···················· | ····················· | ···················· | ···················· | ···················· |
| R | 2110 hCoV-19/Australia/VIC12771/2020\|EPI_ISL_640452\|202 | ···················· | ···················· | ····················· | ···················· | ···················· | ···················· |
| R | 2111 hCoV-19/Australia/VIC12614/2020\|EPI_ISL_640454\|202 | ···················· | ···················· | ····················· | ···················· | ···················· | ···················· |
| R | 2112 hCoV-19/Australia/VIC15944/2020\|EPI_ISL_640455\|202 | ···················· | ···················· | ····················· | ···················· | ···················· | ···················· |
| R | 2113 hCoV-19/Australia/VIC12699/2020\|EPI_ISL_640456\|202 | ···················· | ···················· | ····················· | ···················· | ···················· | ···················· |
| R | 2115 hCoV-19/Australia/VIC16389/2020\|EPI_ISL_640459\|202 | ···················· | ···················· | ····················· | ···················· | ···················· | ···················· |
| R | 2116 hCoV-19/Australia/VIC12711/2020\|EPI_ISL_640460\|202 | ···················· | ···················· | ····················· | ···················· | ···················· | ···················· |
| R | 2117 hCoV-19/Australia/VIC16055/2020\|EPI_ISL_640461\|202 | ···················· | ···················· | ····················· | ···················· | ···················· | ···················· |
| R | 2118 hCoV-19/Australia/VIC12231/2020\|EPI_ISL_640462\|202 | ···················· | ···················· | ····················· | ···················· | ···················· | ···················· |
| R | 2119 hCoV-19/Australia/VIC16034/2020\|EPI_ISL_640463\|202 | ···················· | ···················· | ····················· | ···················· | ···················· | ···················· |
| R | 2120 hCoV-19/Australia/VIC12482/2020\|EPI_ISL_640464\|202 | ···················· | ···················· | ····················· | ···················· | ···················· | ···················· |
| R | 2121 hCoV-19/Australia/VIC12861/2020\|EPI_ISL_640465\|202 | ···················· | ···················· | ····················· | ···················· | ···················· | ···················· |
| R | 2122 hCoV-19/Australia/VIC16307/2020\|EPI_ISL_640466\|202 | ···················· | ···················· | ····················· | ···················· | ···················· | ···················· |
| R | 2123 hCoV-19/Australia/VIC16072/2020\|EPI_ISL_640467\|202 | ···················· | ···················· | ····················· | ···················· | ···················· | ···················· |
| R | 2124 hCoV-19/Australia/VIC16117/2020\|EPI_ISL_640468\|202 | ···················· | ···················· | ····················· | ···················· | ···················· | ···················· |
| R | 2125 hCoV-19/Australia/VIC12455/2020\|EPI_ISL_640469\|202 | ···················· | ···················· | ····················· | ···················· | ···················· | ···················· |
| R | 2126 hCoV-19/Australia/VIC16308/2020\|EPI_ISL_640470\|202 | ···················· | ···················· | ····················· | ···················· | ···················· | ···················· |
| R | 2127 hCoV-19/Australia/VIC16670/2020\|EPI_ISL_640472\|202 | ···················· | ···················· | ····················· | ···················· | ··YK················ | ···················· |
| R | 2128 hCoV-19/Australia/VIC11993/2020\|EPI_ISL_640473\|202 | ···················· | ···················· | ····················· | ···················· | ···················· | ···················· |
| R | 2129 hCoV-19/Australia/VIC16453/2020\|EPI_ISL_640475\|202 | ···················· | ···················· | ····················· | ···················· | ···················· | ···················· |
| R | 2130 hCoV-19/England/QEUH-B13F7B/2020\|EPI_ISL_641687\|20 | ···················· | ···················· | ····················· | ···················· | ···················· | ···················· |
| R | 2131 hCoV-19/England/QEUH-B13DCC/2020\|EPI_ISL_641689\|20 | ···················· | ···················· | ····················· | ···················· | ···················· | ···················· |
| R | 2132 hCoV-19/England/QEUH-B1399B/2020\|EPI_ISL_641690\|20 | ···················· | ···················· | ····················· | ···················· | ···················· | ···················· |
| R | 2133 hCoV-19/England/QEUH-B1395F/2020\|EPI_ISL_641691\|20 | ···················· | ···················· | ····················· | ···················· | ···················· | ···················· |
| R | 2134 hCoV-19/England/QEUH-B13E8D/2020\|EPI_ISL_641692\|20 | ···················· | ···················· | ····················· | ···················· | ···················· | ···················· |
| R | 2135 hCoV-19/England/QEUH-B140A4/2020\|EPI_ISL_641693\|20 | ···················· | ···················· | ····················· | ···················· | ······T············· | ···················· |
| R | 2136 hCoV-19/England/QEUH-B13B1D/2020\|EPI_ISL_641697\|20 | ···················· | ···················· | ····················· | ···················· | ···················· | ···················· |
| R | 2137 hCoV-19/England/QEUH-B1388F/2020\|EPI_ISL_641706\|20 | ···················· | ···················· | ····················· | ···················· | ···················· | ···················· |
| R | 2138 hCoV-19/England/QEUH-B13D35/2020\|EPI_ISL_641710\|20 | ···················· | ···················· | ····················· | ···················· | ···················· | ···················· |
| R | 2139 hCoV-19/England/QEUH-B1355B/2020\|EPI_ISL_641711\|20 | ···················· | ···················· | ····················· | ···················· | ···················· | ···················· |
| R | 2140 hCoV-19/England/QEUH-B10679/2020\|EPI_ISL_641712\|20 | ···················· | ·A·················· | ····················· | ···················· | ···················· | ···················· |
| R | 2141 hCoV-19/England/QEUH-B140FF/2020\|EPI_ISL_641713\|20 | ···················· | ···················· | ····················· | ···················· | ···················· | ···················· |
| R | 2142 hCoV-19/England/QEUH-B133E8/2020\|EPI_ISL_641714\|20 | ···················· | ···················· | ····················· | ···················· | ···················· | ···················· |
| R | 2143 hCoV-19/England/QEUH-B136EF/2020\|EPI_ISL_641715\|20 | ···················· | ···················· | ····················· | ···················· | ···················· | ···················· |
| R | 2144 hCoV-19/England/QEUH-B138E9/2020\|EPI_ISL_641716\|20 | ···················· | ···················· | ····················· | ···················· | ···················· | ···················· |
| R | 2145 hCoV-19/England/QEUH-B13737/2020\|EPI_ISL_641717\|20 | ···················· | ···················· | ····················· | ···················· | ···················· | ···················· |
| R | 2146 hCoV-19/England/QEUH-B138CB/2020\|EPI_ISL_641718\|20 | ···················· | ···················· | ····················· | ···················· | ···················· | ···················· |
| R | 2147 hCoV-19/England/QEUH-B138AD/2020\|EPI_ISL_641719\|20 | ···················· | ···················· | ····················· | ···················· | ···················· | ···················· |
| R | 2148 hCoV-19/England/QEUH-B13782/2020\|EPI_ISL_641720\|20 | ···················· | ···················· | ····················· | ···················· | ···················· | ···················· |
| R | 2149 hCoV-19/England/QEUH-B136FE/2020\|EPI_ISL_641721\|20 | ···················· | ···················· | ····················· | ···················· | ···················· | ···················· |
| R | 2150 hCoV-19/England/QEUH-B13AC5/2020\|EPI_ISL_641722\|20 | ···················· | ···················· | ····················· | ···················· | ···················· | ···················· |
| R | 2151 hCoV-19/England/QEUH-B10785/2020\|EPI_ISL_641723\|20 | ···················· | ·A·················· | ····················· | ···················· | ···················· | ···················· |
| R | 2152 hCoV-19/England/QEUH-B1398C/2020\|EPI_ISL_641724\|20 | ···················· | ···················· | ····················· | ···················· | ···················· | ···················· |
| R | 2153 hCoV-19/England/QEUH-B136C1/2020\|EPI_ISL_641725\|20 | ·····G·············· | ···················· | ····················· | ···················· | ···················· | ···················· |
| R | 2154 hCoV-19/England/QEUH-B13412/2020\|EPI_ISL_641726\|20 | ···················· | ···················· | ····················· | ···················· | ···················· | ···················· |
| R | 2155 hCoV-19/England/QEUH-B12FE5/2020\|EPI_ISL_641727\|20 | ···················· | ···················· | ····················· | ···················· | ···················· | ···················· |
| R | 2156 hCoV-19/England/QEUH-B10600/2020\|EPI_ISL_641728\|20 | ···················· | ···················· | ····················· | ···················· | ···················· | ···················· |
| R | 2157 hCoV-19/England/QEUH-B14569/2020\|EPI_ISL_641729\|20 | ···················· | ···················· | ····················· | ···················· | ···················· | ···················· |
| R | 2158 hCoV-19/England/QEUH-B141A1/2020\|EPI_ISL_641730\|20 | ···················· | ···················· | ····················· | ···················· | ···················· | ···················· |
| R | 2159 hCoV-19/England/QEUH-B13773/2020\|EPI_ISL_641731\|20 | ···················· | ···················· | ····················· | ···················· | ···················· | ···················· |
| R | 2160 hCoV-19/England/QEUH-B14314/2020\|EPI_ISL_641732\|20 | ···················· | ···················· | ····················· | ···················· | ···················· | ···················· |
| R | 2161 hCoV-19/England/QEUH-B13A7A/2020\|EPI_ISL_641733\|20 | ···················· | ···················· | ····················· | ···················· | ···················· | ···················· |
| R | 2162 hCoV-19/England/QEUH-B12F12/2020\|EPI_ISL_641734\|20 | ···················· | ···················· | ····················· | ···················· | ···················· | ···················· |
| R | 2163 hCoV-19/England/QEUH-B13AE3/2020\|EPI_ISL_641735\|20 | ···················· | ···················· | ····················· | ···················· | ···················· | ···················· |
| R | 2164 hCoV-19/England/QEUH-B1063D/2020\|EPI_ISL_641736\|20 | ···················· | ···················· | ····················· | ···················· | ···················· | ···················· |
| R | 2165 hCoV-19/England/QEUH-B141DE/2020\|EPI_ISL_641737\|20 | ···················· | ···················· | ····················· | ···················· | ····T··············· | ···················· |
| R | 2166 hCoV-19/England/QEUH-B0F72F/2020\|EPI_ISL_641739\|20 | ···················· | ···················· | ····················· | ···················· | ···················· | ···················· |
| R | 2167 hCoV-19/England/QEUH-B1080A/2020\|EPI_ISL_641740\|20 | ···················· | ···················· | ····················· | ···················· | ···················· | ···················· |
| R | 2168 hCoV-19/England/QEUH-B1410B/2020\|EPI_ISL_641741\|20 | ···················· | ···················· | ····················· | ···················· | ···················· | ···················· |
| R | 2169 hCoV-19/England/QEUH-B14165/2020\|EPI_ISL_641743\|20 | ···················· | ···················· | ····················· | ···················· | ···················· | ···················· |
| R | 2170 hCoV-19/England/QEUH-B13CCF/2020\|EPI_ISL_641744\|20 | ···················· | ···················· | ····················· | ···················· | ···················· | ···················· |
| R | 2171 hCoV-19/England/QEUH-B1356A/2020\|EPI_ISL_641745\|20 | ···················· | ···················· | ····················· | ···················· | ···················· | ···················· |
| R | 2172 hCoV-19/England/QEUH-B140C2/2020\|EPI_ISL_641746\|20 | ···················· | ···················· | ····················· | ···················· | ···················· | ···················· |
| R | 2173 hCoV-19/England/QEUH-B12F8B/2020\|EPI_ISL_641747\|20 | ···················· | ···················· | ····················· | ···················· | ···················· | ···················· |
| R | 2174 hCoV-19/England/QEUH-B1396E/2020\|EPI_ISL_641748\|20 | ···················· | ···················· | ····················· | ···················· | ···················· | ···················· |
| R | 2175 hCoV-19/England/QEUH-B14305/2020\|EPI_ISL_641749\|20 | ···················· | ···················· | ····················· | ···················· | ···················· | ···················· |
| R | 2176 hCoV-19/England/QEUH-B12C66/2020\|EPI_ISL_641750\|20 | ···················· | ···················· | ····················· | ···················· | ···················· | ···················· |
| R | 2177 hCoV-19/England/QEUH-B13A98/2020\|EPI_ISL_641751\|20 | ···················· | ···················· | ····················· | ···················· | ···················· | ···················· |
| R | 2178 hCoV-19/England/QEUH-B14217/2020\|EPI_ISL_641752\|20 | ···················· | ···················· | ····················· | ···················· | ···················· | ···················· |
| R | 2179 hCoV-19/England/QEUH-B137CE/2020\|EPI_ISL_641753\|20 | ···················· | ···················· | ····················· | ···················· | ···················· | ···················· |
| R | 2180 hCoV-19/England/QEUH-B13EAB/2020\|EPI_ISL_641754\|20 | ···················· | ···················· | ····················· | ···················· | ···················· | ···················· |
| R | 2181 hCoV-19/England/QEUH-B13E23/2020\|EPI_ISL_641755\|20 | ···················· | ···················· | ····················· | ···················· | ···················· | ···················· |
| R | 2182 hCoV-19/England/QEUH-B134F4/2020\|EPI_ISL_641756\|20 | ···················· | ·A·················· | ····················· | ···················· | ···················· | ···A················ |
| R | 2183 hCoV-19/England/QEUH-B12CB1/2020\|EPI_ISL_641758\|20 | ···················· | ···················· | ····················· | ···················· | ···················· | ···················· |
| R | 2184 hCoV-19/England/QEUH-B139C8/2020\|EPI_ISL_641759\|20 | ···················· | ·A·················· | ····················· | ···················· | ···················· | ···················· |
| R | 2185 hCoV-19/England/QEUH-B106E2/2020\|EPI_ISL_641760\|20 | ···················· | ···················· | ····················· | ···················· | ···················· | ···················· |
| R | 2186 hCoV-19/England/QEUH-B13E7E/2020\|EPI_ISL_641762\|20 | ···················· | ···················· | ····················· | ···················· | ···················· | ···················· |

R 2187 hCoV-19/England/QEUH-B12CFD/2020|EPI_ISL_641763|20 ···················· ···················· ···················· ···················· ···················· ····················
R 2188 hCoV-19/England/QEUH-B13579/2020|EPI_ISL_641764|20 ···················· ···················· ···················· ···················· ···················· ····················
R 2189 hCoV-19/England/QEUH-B134A9/2020|EPI_ISL_641765|20 ···················· ·A·················· ···················· ···················· ···················· ····················
R 2190 hCoV-19/England/QEUH-B102C0/2020|EPI_ISL_641767|20 ···················· ···················· ···················· ···················· ···················· ····················
R 2191 hCoV-19/England/QEUH-B13904/2020|EPI_ISL_641768|20 ···················· ···················· ···················· ···················· ···················· ···············A····
R 2192 hCoV-19/England/QEUH-B13746/2020|EPI_ISL_641769|20 ···················· ···················· ···················· ···················· ···················· ····················
R 2193 hCoV-19/England/QEUH-B13843/2020|EPI_ISL_641770|20 ···················· ···················· ···················· ···················· ···················· ····················
R 2194 hCoV-19/England/QEUH-B14077/2020|EPI_ISL_641771|20 ···················· ···················· ···················· ···················· ···················· ····················
R 2195 hCoV-19/England/QEUH-B13500/2020|EPI_ISL_641772|20 ···················· ···················· ···················· ···················· ···················· ····················
R 2196 hCoV-19/England/QEUH-B14095/2020|EPI_ISL_641773|20 ···················· ···················· ···················· ···················· ···················· ····················
R 2197 hCoV-19/England/QEUH-B138DA/2020|EPI_ISL_641774|20 ···················· ···················· ···················· ···················· ···················· ····················
R 2198 hCoV-19/England/QEUH-B13D17/2020|EPI_ISL_641775|20 ···················· ···················· ···················· ···················· ···················· ····················
R 2199 hCoV-19/England/QEUH-B0F83B/2020|EPI_ISL_641776|20 ···················· ···················· ···················· ···················· ···················· ····················
R 2200 hCoV-19/England/QEUH-B1354C/2020|EPI_ISL_641777|20 ···················· ···················· ···················· ···················· ···················· ····················
R 2201 hCoV-19/England/QEUH-B145A5/2020|EPI_ISL_641778|20 ···················· ···················· ···················· ···················· ···················· ····················
R 2202 hCoV-19/England/QEUH-B14156/2020|EPI_ISL_641779|20 ···················· ···················· ···················· ···················· ···················· ····················
R 2203 hCoV-19/England/QEUH-B13D44/2020|EPI_ISL_641780|20 ···················· ···················· ···················· ···················· ···················· ····················
R 2204 hCoV-19/England/QEUH-B1073A/2020|EPI_ISL_641781|20 ···················· ···················· ···················· ···················· ···················· ····················
R 2205 hCoV-19/England/QEUH-B13B0E/2020|EPI_ISL_641782|20 ···················· ···················· ···················· ···················· ···················· ····················
R 2206 hCoV-19/England/QEUH-B13B86/2020|EPI_ISL_641783|20 ···················· ···················· ···················· ···················· ···················· ····················
R 2207 hCoV-19/England/QEUH-B1346D/2020|EPI_ISL_641784|20 ···················· ···················· ···················· ···················· ···················· ····················
R 2208 hCoV-19/England/QEUH-B13EBA/2020|EPI_ISL_641785|20 ···················· ···················· ···················· ···················· ···················· ····················
R 2209 hCoV-19/England/QEUH-B13852/2020|EPI_ISL_641786|20 ···················· ···················· ···················· ···················· ·····T·············· ····················
R 2210 hCoV-19/England/QEUH-B10767/2020|EPI_ISL_641787|20 ···················· ···················· ···················· ···················· ···················· ····················
R 2211 hCoV-19/England/QEUH-B12CDF/2020|EPI_ISL_641788|20 ···················· ···················· ···················· ···················· ···················· ····················
R 2212 hCoV-19/England/QEUH-B1450F/2020|EPI_ISL_641789|20 ···················· ···················· ···················· ···················· ···················· ····················
R 2213 hCoV-19/England/QEUH-B14253/2020|EPI_ISL_641790|20 ···················· ···················· ···················· ···················· ···················· ····················
R 2214 hCoV-19/England/QEUH-B0F74D/2020|EPI_ISL_641791|20 ···················· ···················· ···················· ···················· ···················· ····················
R 2215 hCoV-19/England/QEUH-B14068/2020|EPI_ISL_641793|20 ···················· ···················· ···················· ···················· ···················· ····················
R 2216 hCoV-19/England/QEUH-B1061F/2020|EPI_ISL_641794|20 ···················· ···················· ···················· ···················· ···················· ····················
R 2217 hCoV-19/England/QEUH-B1397D/2020|EPI_ISL_641795|20 ···················· ···················· ···················· ···················· ···················· ····················
R 2218 hCoV-19/England/QEUH-B13069/2020|EPI_ISL_641796|20 ···················· ···················· ···················· ···················· ···················· ····················
R 2219 hCoV-19/England/QEUH-B0F0E8/2020|EPI_ISL_641798|20 ···················· ···················· ···················· ···················· ···················· ····················
R 2220 hCoV-19/England/QEUH-B13AB6/2020|EPI_ISL_641799|20 ···················· ···················· ···················· ···················· ···················· ····················
R 2221 hCoV-19/England/CAMC-B373C0/2020|EPI_ISL_645253|20 ···················· ···················· ···················· ···················· ···················· ····················
R 2222 hCoV-19/England/CAMC-B3708D/2020|EPI_ISL_645254|20 ···················· ···················· ···················· ···················· ···················· ····················
R 2223 hCoV-19/England/CAMC-B3625B/2020|EPI_ISL_645255|20 ···················· ···················· ···················· ···················· ···················· ····················
R 2224 hCoV-19/England/CAMC-B36A17/2020|EPI_ISL_645256|20 ···················· ···················· ···················· ···················· ···················· ····················
R 2225 hCoV-19/England/CAMC-B3674D/2020|EPI_ISL_645257|20 ···················· ···················· ···················· ···················· ···················· ····················
R 2226 hCoV-19/England/CAMC-B367C5/2020|EPI_ISL_645258|20 ···················· ·A·················· ···················· ···················· ···················· ····················
R 2227 hCoV-19/England/CAMC-B363C1/2020|EPI_ISL_645259|20 ···················· ···················· ···················· ···················· ···················· ····················
R 2228 hCoV-19/England/CAMC-B3632B/2020|EPI_ISL_645261|20 ···················· ···················· ···················· ···················· ···················· ····················
R 2229 hCoV-19/England/CAMC-B370BA/2020|EPI_ISL_645262|20 ···················· ···················· ···················· ···················· ···················· ····················
R 2230 hCoV-19/England/MILK-B3D1FD/2020|EPI_ISL_645263|20 ···················· ···················· ···················· ···················· ···················· ····················
R 2231 hCoV-19/England/MILK-B3D6FE/2020|EPI_ISL_645264|20 ···················· ···················· ···················· ···················· ···················· ····················
R 2232 hCoV-19/England/CAMC-B36033/2020|EPI_ISL_645265|20 ···················· ···················· ···················· ···················· ·····T·············· ····················
R 2233 hCoV-19/England/ALDP-B3F085/2020|EPI_ISL_645266|20 ···················· ···················· ···················· ···················· ···················· ····················
R 2234 hCoV-19/England/CAMC-B373FD/2020|EPI_ISL_645267|20 ···················· ···················· ···················· ···················· ···················· ····················
R 2235 hCoV-19/England/CAMC-B3666E/2020|EPI_ISL_645268|20 ···················· ···················· ···················· ···················· ···················· ····················
R 2236 hCoV-19/England/CAMC-B365E9/2020|EPI_ISL_645269|20 ···················· ···················· ···················· ···················· ···················· ····················
R 2237 hCoV-19/England/CAMC-B3690B/2020|EPI_ISL_645270|20 ···················· ·A·················· ···················· ···················· ···················· ····················
R 2238 hCoV-19/England/CAMC-B371C6/2020|EPI_ISL_645271|20 ···················· ···················· ···················· ···················· ···················· ····················
R 2239 hCoV-19/England/CAMC-B35FCE/2020|EPI_ISL_645272|20 ···················· ···················· ···················· ···················· ·····T·············· ····················
R 2240 hCoV-19/England/CAMC-B372A5/2020|EPI_ISL_645273|20 ···················· ···················· ···················· ···················· ·····T·············· ····················
R 2241 hCoV-19/England/CAMC-B35F28/2020|EPI_ISL_645274|20 ···················· ···················· ···················· ···················· ···················· ····················
R 2242 hCoV-19/England/CAMC-B3758E/2020|EPI_ISL_645275|20 ···················· ···················· ···················· ···················· ···················· ····················
R 2243 hCoV-19/England/CAMC-B3676B/2020|EPI_ISL_645276|20 ···················· ···················· ···················· ···················· ···················· ····················
R 2244 hCoV-19/England/CAMC-B35EC1/2020|EPI_ISL_645277|20 ···················· ···················· ···················· ···················· ···················· ····················
R 2245 hCoV-19/England/CAMC-B3675C/2020|EPI_ISL_645278|20 ···················· ···················· ···················· ···················· ···················· ····················
R 2246 hCoV-19/England/CAMC-B35E76/2020|EPI_ISL_645279|20 ···················· ···················· ···················· ···················· ···················· ····················
R 2247 hCoV-19/England/CAMC-B37278/2020|EPI_ISL_645280|20 ···················· ···················· ···················· ···················· ···················· ····················
R 2248 hCoV-19/England/CAMC-B364EC/2020|EPI_ISL_645281|20 ···················· ···················· ···················· ···················· ···················· ····················
R 2249 hCoV-19/England/CAMC-B35FA0/2020|EPI_ISL_645283|20 ···················· ···················· ···················· ···················· ···················· ····················
R 2250 hCoV-19/England/CAMC-B369B0/2020|EPI_ISL_645284|20 ···················· ···················· ···················· ···················· ···················· ····················
R 2251 hCoV-19/England/MILK-B3D263/2020|EPI_ISL_645285|20 ···················· ···················· ···················· ···················· ···················· ····················
R 2252 hCoV-19/England/CAMC-B3714E/2020|EPI_ISL_645286|20 ···················· ···················· ···················· ···················· ···················· ····················
R 2253 hCoV-19/England/CAMC-B37506/2020|EPI_ISL_645287|20 ···················· ···················· ···················· ···················· ···················· ····················
R 2254 hCoV-19/England/CAMC-B36428/2020|EPI_ISL_645288|20 ···················· ···················· ···················· ···················· ···················· ····················
R 2255 hCoV-19/England/CAMC-B3707E/2020|EPI_ISL_645289|20 ···················· ···················· ···················· ···················· ·····T·············· ···············A····
R 2256 hCoV-19/England/CAMC-B36473/2020|EPI_ISL_645290|20 ···················· ···················· ···················· ···················· ···················· ····················
R 2257 hCoV-19/England/CAMC-B365CB/2020|EPI_ISL_645291|20 ···················· ···················· ···················· ···················· ·····T·············· ····················
R 2258 hCoV-19/England/CAMC-B35F0A/2020|EPI_ISL_645292|20 ···················· ···················· ···················· ···················· ···················· ····················
R 2259 hCoV-19/England/CAMC-B36FFA/2020|EPI_ISL_645293|20 ···················· ···················· ···················· ···················· ···················· ····················
R 2260 hCoV-19/England/CAMC-B3614F/2020|EPI_ISL_645294|20 ···················· ·A·······A·········· ···················· ···················· ···················· ····················
R 2261 hCoV-19/England/CAMC-B37287/2020|EPI_ISL_645295|20 ···················· ···················· ···················· ···················· ·····T·············· ····················
R 2262 hCoV-19/England/ALDP-B3E8E8/2020|EPI_ISL_645297|20 ···················· ···················· ···················· ···················· ···················· ····················
R 2263 hCoV-19/England/CAMC-B369DE/2020|EPI_ISL_645298|20 ···················· ···················· ···················· ···················· ···················· ····················
R 2264 hCoV-19/England/ALDP-B24CE9/2020|EPI_ISL_646580|20 ···················· ···················· ···················· ···················· ···················· ····················
R 2265 hCoV-19/England/ALDP-B25FEF/2020|EPI_ISL_646581|20 ···················· ···················· ···················· ···················· ···················· ····················
R 2266 hCoV-19/England/QEUH-B2B8A2/2020|EPI_ISL_646582|20 ···················· ···················· ···················· ···················· ···················· ····················

| | | | | | | | |
|---|---|---|---|---|---|---|---|
| R | 2267 hCoV-19/England/ALDP-B263F1/2020\|EPI_ISL_646583\|20 | ···················· | ···················· | ····················· | ···················· | ···················· | ···················· |
| R | 2268 hCoV-19/England/QEUH-B2B866/2020\|EPI_ISL_646584\|20 | ···················· | ···················· | ····················· | ···················· | ···················· | ···················· |
| R | 2269 hCoV-19/England/ALDP-B252C8/2020\|EPI_ISL_646585\|20 | ···················· | ···················· | ····················· | ···················· | ···················· | ···················· |
| R | 2270 hCoV-19/England/ALDP-B2625E/2020\|EPI_ISL_646586\|20 | ···················· | ···················· | ····················· | ···················· | ···················· | ···················· |
| R | 2271 hCoV-19/England/QEUH-B2C971/2020\|EPI_ISL_646588\|20 | ···················· | ···················· | ····················· | ···················· | ···················· | ···················· |
| R | 2272 hCoV-19/England/QEUH-B2BE46/2020\|EPI_ISL_646590\|20 | ···················· | ·A·················· | ····················· | ···················· | ···················· | ···················· |
| R | 2273 hCoV-19/England/ALDP-B24F1D/2020\|EPI_ISL_646591\|20 | ···················· | ···················· | ····················· | ···················· | ···················· | ···················· |
| R | 2274 hCoV-19/England/ALDP-B263E2/2020\|EPI_ISL_646592\|20 | ···················· | ···················· | ····················· | ···················· | ···················· | ···················· |
| R | 2275 hCoV-19/England/QEUH-B2B75A/2020\|EPI_ISL_646593\|20 | ···················· | ···················· | ····················· | ···················· | ······T············· | ···················· |
| R | 2276 hCoV-19/England/ALDP-B24D8C/2020\|EPI_ISL_646594\|20 | ···················· | ···················· | ····················· | ···················· | ···················· | ···················· |
| R | 2277 hCoV-19/England/ALDP-B2527D/2020\|EPI_ISL_646595\|20 | ···················· | ···················· | ····················· | ···················· | ···················· | ···················· |
| R | 2278 hCoV-19/England/QEUH-B2B59C/2020\|EPI_ISL_646596\|20 | ···················· | ···················· | ····················· | ···················· | ···················· | ···················· |
| R | 2279 hCoV-19/England/ALDP-B2518F/2020\|EPI_ISL_646597\|20 | ···················· | ·A·················· | ····················· | ···················· | ···················· | ···················· |
| R | 2280 hCoV-19/England/QEUH-B2CAAB/2020\|EPI_ISL_646598\|20 | ···················· | ···················· | ····················· | ···················· | ···················· | ···················· |
| R | 2281 hCoV-19/England/ALDP-B251E9/2020\|EPI_ISL_646599\|20 | ···················· | ···················· | ····················· | ···················· | ···················· | ···················· |
| R | 2282 hCoV-19/Wales/QEUH-B13F02/2020\|EPI_ISL_647800\|2020 | ···················· | ···················· | ····················· | ···················· | ···················· | ···················· |
| R | 2283 hCoV-19/Wales/QEUH-B13FB7/2020\|EPI_ISL_647801\|2020 | ···················· | ···················· | ····················· | ···················· | ···················· | ···················· |
| R | 2284 hCoV-19/Wales/QEUH-B13825/2020\|EPI_ISL_647802\|2020 | ···················· | ···················· | ····················· | ···················· | ···················· | ···················· |
| R | 2285 hCoV-19/Wales/QEUH-B0DE86/2020\|EPI_ISL_647803\|2020 | ···················· | ···················· | ····················· | ···················· | ···················· | ···················· |
| R | 2286 hCoV-19/Wales/QEUH-B0DF1A/2020\|EPI_ISL_647804\|2020 | ···················· | ···················· | ····················· | ···················· | ···················· | ···················· |
| R | 2287 hCoV-19/Wales/QEUH-B10FBA/2020\|EPI_ISL_647805\|2020 | ···················· | ···················· | ····················· | ···················· | ···················· | ···················· |
| R | 2288 hCoV-19/Wales/QEUH-B0DFDE/2020\|EPI_ISL_647806\|2020 | ···················· | ···················· | ····················· | ···················· | ···················· | ···················· |
| R | 2289 hCoV-19/Wales/MILK-B02BA8/2020\|EPI_ISL_647807\|2020 | ···················· | ···················· | ····················· | ···················· | ···················· | ···················· |
| R | 2290 hCoV-19/Wales/MILK-B02B7B/2020\|EPI_ISL_647808\|2020 | ···················· | ···················· | ····················· | ···················· | ···················· | ···················· |
| R | 2291 hCoV-19/Wales/MILK-B025E6/2020\|EPI_ISL_647809\|2020 | ···················· | ···················· | ····················· | ···················· | ···················· | ···················· |
| R | 2292 hCoV-19/Wales/MILK-B02504/2020\|EPI_ISL_647810\|2020 | ···················· | ···················· | ····················· | ···················· | ···················· | ···················· |
| R | 2293 hCoV-19/Wales/MILK-B02CA5/2020\|EPI_ISL_647812\|2020 | ···················· | ···················· | ····················· | ···················· | ···················· | ···················· |
| R | 2294 hCoV-19/Wales/MILK-B01CF1/2020\|EPI_ISL_647813\|2020 | ···················· | ···················· | ····················· | ···················· | ···················· | ···················· |
| R | 2295 hCoV-19/Wales/MILK-B024BC/2020\|EPI_ISL_647814\|2020 | ···················· | ···················· | ····················· | ···················· | ···················· | ···················· |
| R | 2296 hCoV-19/Wales/MILK-B02B99/2020\|EPI_ISL_647815\|2020 | ···················· | ···················· | ····················· | ···················· | ···················· | ···················· |
| R | 2297 hCoV-19/Wales/MILK-B025C8/2020\|EPI_ISL_647816\|2020 | ···················· | ···················· | ····················· | ···················· | ···················· | ···················· |
| R | 2298 hCoV-19/Wales/MILK-B02D39/2020\|EPI_ISL_647817\|2020 | ···················· | ···················· | ····················· | ···················· | ···················· | ···················· |
| R | 2299 hCoV-19/Wales/MILK-B0259B/2020\|EPI_ISL_647818\|2020 | ···················· | ···················· | ····················· | ···················· | ···················· | ···················· |
| R | 2300 hCoV-19/Wales/MILK-B0639D/2020\|EPI_ISL_647820\|2020 | ···················· | ···················· | ····················· | ···················· | ···················· | ···················· |
| R | 2301 hCoV-19/Wales/MILK-B02D48/2020\|EPI_ISL_647821\|2020 | ···················· | ···················· | ····················· | ···················· | ···················· | ···················· |
| R | 2302 hCoV-19/Wales/MILK-B02A23/2020\|EPI_ISL_647822\|2020 | ···················· | ···················· | ····················· | ···················· | ···················· | ···················· |
| R | 2303 hCoV-19/Wales/MILK-B06236/2020\|EPI_ISL_647823\|2020 | ···················· | ···················· | ····················· | ···················· | ···················· | ···················· |
| R | 2304 hCoV-19/Wales/MILK-B02B02/2020\|EPI_ISL_647824\|2020 | ···················· | ···················· | ····················· | ···················· | ···················· | ···················· |
| R | 2305 hCoV-19/Australia/VIC12446/2020\|EPI_ISL_640363\|202 | ···················· | ···················· | ····················· | ···················· | ···················· | ···················· |
| R | 2307 hCoV-19/Australia/VIC12229/2020\|EPI_ISL_640367\|202 | ···················· | ···················· | ····················· | ···················· | ···················· | ···················· |
| R | 2308 hCoV-19/Australia/VIC16402/2020\|EPI_ISL_640368\|202 | ···················· | ···················· | ····················· | ···················· | ···················· | ···················· |
| R | 2309 hCoV-19/Australia/VIC12432/2020\|EPI_ISL_640369\|202 | ···················· | ···················· | ····················· | ···················· | ···················· | ···················· |
| R | 2310 hCoV-19/Australia/VIC16050/2020\|EPI_ISL_640371\|202 | ···················· | ···················· | ····················· | ···················· | ···················· | ···················· |
| R | 2311 hCoV-19/Australia/VIC12781/2020\|EPI_ISL_640372\|202 | ···················· | ···················· | ····················· | ···················· | ···················· | ···················· |
| R | 2312 hCoV-19/Australia/VIC16313/2020\|EPI_ISL_640373\|202 | ···················· | ···················· | ····················· | ···················· | ···················· | ···················· |
| R | 2313 hCoV-19/Australia/VIC16003/2020\|EPI_ISL_640376\|202 | ···················· | ···················· | ····················· | ···················· | ···················· | ···················· |
| R | 2314 hCoV-19/Australia/VIC12823/2020\|EPI_ISL_640377\|202 | ···················· | ···················· | ····················· | ···················· | ···················· | ···················· |
| R | 2315 hCoV-19/England/QEUH-B106F1/2020\|EPI_ISL_641694\|20 | ···················· | ···················· | ····················· | ···················· | ···················· | ···················· |
| R | 2316 hCoV-19/England/QEUH-B1404A/2020\|EPI_ISL_641695\|20 | ···················· | ···················· | ····················· | ···················· | ···················· | ···················· |
| R | 2317 hCoV-19/England/QEUH-B13E32/2020\|EPI_ISL_641696\|20 | ···················· | ···················· | ····················· | ···················· | ···················· | ···················· |
| R | 2318 hCoV-19/England/QEUH-B10169/2020\|EPI_ISL_641698\|20 | ···················· | ···················· | ····················· | ···················· | ···················· | ···················· |
| R | 2319 hCoV-19/England/QEUH-B134D6/2020\|EPI_ISL_641699\|20 | ···················· | ···················· | ····················· | ···················· | ···················· | ···················· |
| R | 2320 hCoV-19/England/MILK-B01ECE/2020\|EPI_ISL_642901\|20 | ···················· | ·A·················· | ····················· | ···················· | ···················· | ···A················ |
| R | 2321 hCoV-19/England/MILK-B02461/2020\|EPI_ISL_642902\|20 | ···················· | ···················· | ····················· | ···················· | ···················· | ···················· |
| R | 2322 hCoV-19/England/MILK-B01DC1/2020\|EPI_ISL_642904\|20 | ···················· | ···················· | ····················· | ···················· | ···················· | ···················· |
| R | 2323 hCoV-19/England/MILK-B02C0F/2020\|EPI_ISL_642905\|20 | ···················· | ···················· | ····················· | ···················· | ···················· | ···················· |
| R | 2324 hCoV-19/England/MILK-B05FF4/2020\|EPI_ISL_642906\|20 | ···················· | ···················· | ····················· | ···················· | ···················· | ···················· |
| R | 2325 hCoV-19/England/MILK-B05EE8/2020\|EPI_ISL_642907\|20 | ···················· | ···················· | ····················· | ···················· | ···················· | ···················· |
| R | 2326 hCoV-19/England/MILK-B02944/2020\|EPI_ISL_642908\|20 | ···················· | ···················· | ····················· | ···················· | ···················· | ···················· |
| R | 2327 hCoV-19/England/MILK-B02267/2020\|EPI_ISL_642909\|20 | ···················· | ···················· | ····················· | ···················· | ···················· | ···················· |
| R | 2328 hCoV-19/England/MILK-B05E60/2020\|EPI_ISL_642910\|20 | ···················· | ···················· | ····················· | ···················· | ···················· | ···················· |
| R | 2329 hCoV-19/England/MILK-B02698/2020\|EPI_ISL_642911\|20 | ···················· | ···················· | ····················· | ···················· | ···················· | ···················· |
| R | 2330 hCoV-19/England/MILK-B05FC7/2020\|EPI_ISL_642913\|20 | ···················· | ···················· | ····················· | ···················· | ···················· | ···················· |
| R | 2331 hCoV-19/England/MILK-B02C1E/2020\|EPI_ISL_642914\|20 | ···················· | ···················· | ····················· | ···················· | ···················· | ···················· |
| R | 2332 hCoV-19/England/MILK-B0306C/2020\|EPI_ISL_642915\|20 | ···················· | ···················· | ····················· | ···················· | ···················· | ···················· |
| R | 2333 hCoV-19/England/MILK-B05F30/2020\|EPI_ISL_642916\|20 | ···················· | ···················· | ····················· | ···················· | ···················· | ···················· |
| R | 2334 hCoV-19/England/MILK-B031D2/2020\|EPI_ISL_642917\|20 | ···················· | ···················· | ····················· | ···················· | ······T············· | ···················· |
| R | 2335 hCoV-19/England/MILK-B023DD/2020\|EPI_ISL_642918\|20 | ···················· | ···················· | ····················· | ···················· | ···················· | ···················· |
| R | 2336 hCoV-19/England/MILK-B0221C/2020\|EPI_ISL_642919\|20 | ···················· | ·A·················· | ····················· | ···················· | ···················· | ···A················ |
| R | 2337 hCoV-19/England/MILK-B01E28/2020\|EPI_ISL_642920\|20 | ···················· | ···················· | ····················· | ···················· | ···················· | ···················· |
| R | 2338 hCoV-19/England/MILK-B02443/2020\|EPI_ISL_642921\|20 | ···················· | ···················· | ····················· | ···················· | ······T············· | ···················· |
| R | 2339 hCoV-19/England/MILK-B0610C/2020\|EPI_ISL_642922\|20 | ···················· | ···················· | ····················· | ···················· | ···················· | ···················· |
| R | 2340 hCoV-19/England/MILK-B01E37/2020\|EPI_ISL_642923\|20 | ···················· | ···················· | ····················· | ···················· | ···················· | ···················· |
| R | 2341 hCoV-19/England/MILK-B0280B/2020\|EPI_ISL_642924\|20 | ···················· | ···················· | ····················· | ···················· | ···················· | ···················· |
| R | 2342 hCoV-19/England/MILK-B02003/2020\|EPI_ISL_642925\|20 | ···················· | ···················· | ····················· | ···················· | ······T············· | ···················· |
| R | 2343 hCoV-19/England/MILK-B031C3/2020\|EPI_ISL_642926\|20 | ···················· | ···················· | ····················· | ···················· | ···················· | ···················· |
| R | 2344 hCoV-19/England/MILK-B062AF/2020\|EPI_ISL_642927\|20 | ···················· | ···················· | ····················· | ···················· | ···················· | ···················· |
| R | 2345 hCoV-19/England/MILK-B025B9/2020\|EPI_ISL_642928\|20 | ···················· | ···················· | ····················· | ···················· | ···················· | ···················· |
| R | 2346 hCoV-19/France/NAQ-SC219/2020\|EPI_ISL_645170\|2020- | ···················· | ···················· | ····················· | ···················· | ···················· | ···················· |
| R | 2347 hCoV-19/France/NAQ-SC220/2020\|EPI_ISL_645171\|2020- | ···················· | ···················· | ····················· | ···················· | ···················· | ···················· |

| | | | | | | | | |
|---|---|---|---|---|---|---|---|---|
| R | 2348 hCoV-19/France/NAQ-SC231/2020\|EPI_ISL_645172\|2020- | .................... | .................... | .................... | .................... | .................... | .................... | |
| R | 2349 hCoV-19/France/PAC-SC233/2020\|EPI_ISL_645173\|2020- | .................... | .................... | .................... | .................... | .................... | .................... | |
| R | 2350 hCoV-19/France/NAQ-SC251/2020\|EPI_ISL_645174\|2020- | .................... | .................... | .................... | .................... | .................... | .................... | |
| R | 2351 hCoV-19/France/NAQ-SC254/2020\|EPI_ISL_645175\|2020- | .................... | .................... | .................... | .................... | .................... | .................... | |
| R | 2352 hCoV-19/France/NAQ-SC255/2020\|EPI_ISL_645176\|2020- | .................... | .................... | .................... | .................... | .................... | .................... | |
| R | 2353 hCoV-19/France/NAQ-SC256/2020\|EPI_ISL_645177\|2020- | .................... | .................... | .................... | .................... | .................... | .................... | |
| R | 2354 hCoV-19/France/NAQ-SC258/2020\|EPI_ISL_645178\|2020- | .................... | .................... | .................... | .................... | .................... | .................... | |
| R | 2355 hCoV-19/France/NAQ-SC273/2020\|EPI_ISL_645179\|2020- | .................... | .................... | .................... | .................... | .................... | .................... | |
| R | 2356 hCoV-19/France/NAQ-SC274/2020\|EPI_ISL_645180\|2020- | .................... | .................... | .................... | .................... | .................... | .................... | |
| R | 2357 hCoV-19/France/NAQ-SC277/2020\|EPI_ISL_645181\|2020- | .................... | .................... | .................... | .................... | .................... | .................... | |
| R | 2358 hCoV-19/France/NAQ-SC278/2020\|EPI_ISL_645182\|2020- | .................... | .................... | .................... | .................... | .................... | .................... | |
| R | 2359 hCoV-19/France/NAQ-SC279/2020\|EPI_ISL_645183\|2020- | .................... | .................... | .................... | .................... | .................... | .................... | |
| R | 2360 hCoV-19/France/ARA-SC364/2020\|EPI_ISL_645184\|2020- | .................... | .................... | .................... | .................... | .................... | .................... | |
| R | 2361 hCoV-19/France/ARA-SC369/2020\|EPI_ISL_645185\|2020- | .................... | .................... | .................... | .................... | .................... | .................... | |
| R | 2362 hCoV-19/France/ARA-SC373/2020\|EPI_ISL_645186\|2020- | .................... | .................... | .................... | .................... | .................... | .................... | |
| R | 2363 hCoV-19/France/ARA-SC376/2020\|EPI_ISL_645187\|2020- | .................... | .................... | .................... | .................... | .................... | .................... | |
| R | 2364 hCoV-19/France/ARA-SC377/2020\|EPI_ISL_645188\|2020- | .................... | .................... | .................... | .................... | .................... | .................... | |
| R | 2365 hCoV-19/France/ARA-SC379/2020\|EPI_ISL_645189\|2020- | .................... | .................... | .................... | .................... | .................... | .................... | |
| R | 2366 hCoV-19/France/ARA-SC383/2020\|EPI_ISL_645190\|2020- | .................... | .................... | .................... | .................... | .................... | .................... | |
| R | 2367 hCoV-19/France/ARA-SC387/2020\|EPI_ISL_645191\|2020- | .................... | .................... | .................... | .................... | .................... | .................... | |
| R | 2368 hCoV-19/France/ARA-SC388/2020\|EPI_ISL_645192\|2020- | .................... | .................... | .................... | .................... | .................... | .................... | |
| R | 2369 hCoV-19/France/ARA-SC390/2020\|EPI_ISL_645193\|2020- | .................... | .................... | .................... | .................... | .................... | .................... | |
| R | 2370 hCoV-19/France/ARA-SC392/2020\|EPI_ISL_645194\|2020- | .................... | .................... | .................... | .................... | .................... | .................... | |
| R | 2371 hCoV-19/France/ARA-SC394/2020\|EPI_ISL_645195\|2020- | .................... | .................... | .................... | .................... | .................... | .................... | |
| R | 2372 hCoV-19/France/ARA-SC395/2020\|EPI_ISL_645196\|2020- | .................... | .................... | .................... | .................... | .................... | .................... | |
| R | 2373 hCoV-19/France/OCC-SC534/2020\|EPI_ISL_645197\|2020- | .................... | ...................A | .................... | .................... | .................... | .................... | 1 |
| R | 2374 hCoV-19/France/OCC-SC535/2020\|EPI_ISL_645198\|2020- | .................... | .................... | .................... | .................... | .................... | .................... | |
| R | 2375 hCoV-19/France/OCC-SC540/2020\|EPI_ISL_645199\|2020- | .................... | .................... | .................... | .................... | .................... | .................... | |
| R | 2376 hCoV-19/France/OCC-SC543/2020\|EPI_ISL_645200\|2020- | .................... | .A.................. | .................... | .................... | .................... | .................... | |
| R | 2377 hCoV-19/France/OCC-SC545/2020\|EPI_ISL_645201\|2020- | .................... | .................... | .................... | .................... | .................... | .................... | |
| R | 2378 hCoV-19/France/OCC-SC546/2020\|EPI_ISL_645202\|2020- | .................... | .................... | .................... | .................... | .................... | .................... | |
| R | 2379 hCoV-19/France/OCC-SC547/2020\|EPI_ISL_645203\|2020- | .................... | .................... | .................... | .................... | .................... | .................... | |
| R | 2380 hCoV-19/France/OCC-SC549/2020\|EPI_ISL_645204\|2020- | .................... | ..........A......... | .................... | .................... | .................... | .................... | |
| R | 2381 hCoV-19/France/OCC-SC553/2020\|EPI_ISL_645205\|2020- | .................... | ..........A......... | .................... | .................... | .................... | .................... | |
| R | 2382 hCoV-19/France/OCC-SC554/2020\|EPI_ISL_645206\|2020- | .................... | .................... | .................... | .................... | .................... | .................... | |
| R | 2383 hCoV-19/France/OCC-SC559/2020\|EPI_ISL_645207\|2020- | .................... | .................... | .................... | .................... | .................... | .................... | |
| R | 2384 hCoV-19/France/OCC-SC562/2020\|EPI_ISL_645208\|2020- | .................... | ..........A......... | .................... | .................... | .................... | .................... | |
| R | 2385 hCoV-19/France/OCC-SC572/2020\|EPI_ISL_645209\|2020- | .................... | ..........A......... | .................... | .................... | .................... | .................... | |
| R | 2386 hCoV-19/France/PAC-SC456/2020\|EPI_ISL_645212\|2020- | .................... | .................... | .................... | .................... | .................... | .................... | |
| R | 2387 hCoV-19/France/PAC-SC460/2020\|EPI_ISL_645213\|2020- | .................... | .................... | .................... | .................... | .................... | .................... | |
| R | 2388 hCoV-19/France/PAC-SC461/2020\|EPI_ISL_645214\|2020- | .................... | .................... | .................... | .................... | .................... | .................... | |
| R | 2389 hCoV-19/France/OCC-SC574/2020\|EPI_ISL_645215\|2020- | .................... | .................... | ...........T........ | .................... | .................... | .................... | |
| R | 2390 hCoV-19/France/NAQ-SC575/2020\|EPI_ISL_645216\|2020- | .................... | .................... | .................... | .................... | .................... | .................... | |
| R | 2391 hCoV-19/France/PDL-SC601/2020\|EPI_ISL_645217\|2020- | .................... | .................... | .................... | .................... | .................... | .................... | |
| R | 2392 hCoV-19/France/PDL-SC649/2020\|EPI_ISL_645218\|2020- | .................... | .................... | .................... | .................... | .................... | .................... | |
| R | 2393 hCoV-19/England/CAMC-B36042/2020\|EPI_ISL_645219\|20 | .................... | .................... | .................... | .................... | .................... | .................... | |
| R | 2394 hCoV-19/England/CAMC-B35EFE/2020\|EPI_ISL_645220\|20 | .................... | .................... | .................... | .................... | .................... | .................... | |
| R | 2395 hCoV-19/England/MILK-B3D56A/2020\|EPI_ISL_645221\|20 | .................... | .................... | .................... | .................... | ......T............. | .................... | |
| R | 2396 hCoV-19/England/CAMC-B36446/2020\|EPI_ISL_645222\|20 | .................... | .................... | .................... | .................... | .................... | .................... | |
| R | 2397 hCoV-19/England/CAMC-B360CA/2020\|EPI_ISL_645223\|20 | .................... | .................... | .................... | .................... | .................... | .................... | |
| R | 2398 hCoV-19/England/CAMC-B370C9/2020\|EPI_ISL_645224\|20 | .................... | .................... | .................... | .................... | .................... | .................... | |
| R | 2399 hCoV-19/England/CAMC-B36956/2020\|EPI_ISL_645225\|20 | .................... | .................... | .................... | .................... | .................... | .................... | |
| R | 2400 hCoV-19/England/MILK-B3D49A/2020\|EPI_ISL_645226\|20 | .................... | .................... | .................... | .................... | .................... | .................... | |
| R | 2401 hCoV-19/England/CAMC-B363A3/2020\|EPI_ISL_645228\|20 | .................... | .................... | .................... | .................... | .................... | .................... | |
| R | 2402 hCoV-19/England/CAMC-B36A44/2020\|EPI_ISL_645229\|20 | .................... | .................... | .................... | .................... | .................... | .................... | |
| R | 2403 hCoV-19/England/CAMC-B36455/2020\|EPI_ISL_645230\|20 | .................... | .A.......A.......... | .................... | .................... | .................... | .................... | |
| R | 2404 hCoV-19/England/CAMC-B3724B/2020\|EPI_ISL_645231\|20 | .................... | .................... | .................... | .................... | .................... | .................... | |
| R | 2405 hCoV-19/England/CAMC-B37032/2020\|EPI_ISL_645232\|20 | .................... | .A.................. | .................... | .................... | .................... | ...A................ | |
| R | 2406 hCoV-19/England/CAMC-B35F64/2020\|EPI_ISL_645234\|20 | .................... | .................... | .................... | .................... | .................... | .................... | |
| R | 2407 hCoV-19/England/CAMC-B36349/2020\|EPI_ISL_645235\|20 | .................... | .................... | .................... | .................... | ......T............. | .................... | |
| R | 2408 hCoV-19/England/CAMC-B37375/2020\|EPI_ISL_645236\|20 | .................... | .................... | .................... | .................... | .................... | .................... | |
| R | 2409 hCoV-19/England/CAMC-B373A2/2020\|EPI_ISL_645237\|20 | .................... | .................... | .................... | .................... | .................... | .................... | |
| R | 2410 hCoV-19/England/CAMC-B36A26/2020\|EPI_ISL_645238\|20 | .................... | .................... | .................... | .................... | .................... | .................... | |
| R | 2411 hCoV-19/England/CAMC-B373B1/2020\|EPI_ISL_645239\|20 | .................... | .................... | .................... | .................... | .................... | .................... | |
| R | 2412 hCoV-19/England/ALDP-B3E736/2020\|EPI_ISL_645240\|20 | .................... | .................... | .................... | .................... | .................... | .................... | |
| R | 2413 hCoV-19/England/MILK-B3D1EE/2020\|EPI_ISL_645241\|20 | .................... | .................... | .................... | .................... | .................... | .................... | |
| R | 2414 hCoV-19/England/CAMC-B367D4/2020\|EPI_ISL_645242\|20 | .................... | .................... | .................... | .................... | .................... | .................... | |
| R | 2415 hCoV-19/England/MILK-B3D209/2020\|EPI_ISL_645244\|20 | .................... | .................... | .................... | .................... | .................... | .................... | |
| R | 2416 hCoV-19/England/CAMC-B367F2/2020\|EPI_ISL_645245\|20 | .................... | .................... | .................... | .................... | ......T............. | .................... | |
| R | 2417 hCoV-19/England/CAMC-B36983/2020\|EPI_ISL_645247\|20 | .................... | .................... | .................... | .................... | .................... | .................... | |
| R | 2418 hCoV-19/England/CAMC-B370AB/2020\|EPI_ISL_645249\|20 | .................... | .................... | .................... | .................... | .................... | .................... | |
| R | 2419 hCoV-19/England/QEUH-B2B435/2020\|EPI_ISL_646482\|20 | .................... | .................... | .................... | .................... | .................... | .................... | |
| R | 2420 hCoV-19/England/QEUH-B2B462/2020\|EPI_ISL_646483\|20 | .................... | .................... | .................... | .................... | .................... | .................... | |
| R | 2421 hCoV-19/England/QEUH-B2BA60/2020\|EPI_ISL_646484\|20 | .................... | .................... | .................... | .................... | .................... | .................... | |
| R | 2422 hCoV-19/England/ALDP-B250A0/2020\|EPI_ISL_646485\|20 | .................... | .................... | .................... | .................... | .................... | .................... | |
| R | 2423 hCoV-19/England/ALDP-B26494/2020\|EPI_ISL_646500\|20 | .................... | .................... | .................... | .................... | .................... | .................... | |
| R | 2424 hCoV-19/England/QEUH-B2B875/2020\|EPI_ISL_646501\|20 | .................... | .................... | .................... | .................... | .................... | .................... | |
| R | 2425 hCoV-19/England/ALDP-B26212/2020\|EPI_ISL_646502\|20 | .................... | .................... | .................... | .................... | .................... | .................... | |
| R | 2426 hCoV-19/England/QEUH-B2B6F3/2020\|EPI_ISL_646503\|20 | .................... | .................... | .................... | .................... | .................... | .................... | |
| R | 2427 hCoV-19/England/QEUH-B2B787/2020\|EPI_ISL_646504\|20 | .................... | .................... | .................... | .................... | .................... | .................... | |

| | | | | | | | | |
|---|---|---|---|---|---|---|---|---|
| R | 2428 | hCoV-19/England/ALDP-B25F58/2020\|EPI_ISL_646505\|20 | ···················· | ···················· | ···················· | ···················· | ···················· | ···················· |
| R | 2429 | hCoV-19/England/ALDP-B25213/2020\|EPI_ISL_646506\|20 | ···················· | ···················· | ···················· | ···················· | ···················· | ···················· |
| R | 2430 | hCoV-19/England/QEUH-B2B65D/2020\|EPI_ISL_646507\|20 | ···················· | ···················· | ···················· | ···················· | ···················· | ···················· |
| R | 2431 | hCoV-19/England/ALDP-B261E8/2020\|EPI_ISL_646508\|20 | ···················· | ···A················ | ···················· | ···················· | ···················· | ···················· |
| R | 2432 | hCoV-19/England/ALDP-B24D5F/2020\|EPI_ISL_646509\|20 | ···················· | ·A·················· | ···················· | ···················· | ···················· | ···················· |
| R | 2433 | hCoV-19/England/ALDP-B24EA7/2020\|EPI_ISL_646511\|20 | ···················· | ···················· | ···················· | ···················· | ···················· | ···················· |
| R | 2434 | hCoV-19/England/ALDP-B2642B/2020\|EPI_ISL_646512\|20 | ···················· | ·A·················· | ···················· | ···················· | ···················· | ···················· |
| R | 2435 | hCoV-19/England/ALDP-B25152/2020\|EPI_ISL_646514\|20 | ···················· | ···················· | ···················· | ···················· | ···················· | ···················· |
| R | 2436 | hCoV-19/England/ALDP-B25028/2020\|EPI_ISL_646515\|20 | ···················· | ···················· | ···················· | ···················· | ···················· | ···················· |
| R | 2437 | hCoV-19/England/ALDP-B251DA/2020\|EPI_ISL_646516\|20 | ···················· | ···················· | ···················· | ···················· | ···················· | ···················· |
| R | 2438 | hCoV-19/England/QEUH-B2C892/2020\|EPI_ISL_646517\|20 | ···················· | ···················· | ···················· | ···················· | ···················· | ···················· |
| R | 2439 | hCoV-19/England/ALDP-B251CB/2020\|EPI_ISL_646518\|20 | ···················· | ···················· | ···················· | ···················· | ···················· | ···················· |
| R | 2440 | hCoV-19/England/QEUH-B2B5AB/2020\|EPI_ISL_646519\|20 | ···················· | ···················· | ···················· | ···················· | ···················· | ···················· |
| R | 2441 | hCoV-19/England/ALDP-B261BB/2020\|EPI_ISL_646520\|20 | ···················· | ···················· | ···················· | ···················· | ···················· | ···················· |
| R | 2442 | hCoV-19/England/ALDP-B25F67/2020\|EPI_ISL_646521\|20 | ···················· | ···················· | ···················· | ···················· | ···················· | ···················· |
| R | 2443 | hCoV-19/England/ALDP-B25F3A/2020\|EPI_ISL_646522\|20 | ···················· | ···················· | ···················· | ···················· | ···················· | ···················· |
| R | 2444 | hCoV-19/England/QEUH-B2CA7E/2020\|EPI_ISL_646523\|20 | ···················· | ·A·················· | ···················· | ···················· | ···················· | ··A················· |
| R | 2445 | hCoV-19/England/QEUH-B2B329/2020\|EPI_ISL_646524\|20 | ···················· | ···················· | ···················· | ···················· | ···················· | ···················· |
| R | 2446 | hCoV-19/England/ALDP-B24E7A/2020\|EPI_ISL_646526\|20 | ···················· | ···················· | ···················· | ···················· | ···················· | ···················· |
| R | 2447 | hCoV-19/England/QEUH-B2B918/2020\|EPI_ISL_646527\|20 | ···················· | ···················· | ···················· | ···················· | ···················· | ···················· |
| R | 2448 | hCoV-19/England/ALDP-B25046/2020\|EPI_ISL_646528\|20 | ···················· | ···················· | ···················· | ···················· | ···················· | ···················· |
| R | 2449 | hCoV-19/England/ALDP-B25116/2020\|EPI_ISL_646529\|20 | ···················· | ···················· | ···················· | ···················· | ···················· | ···················· |
| R | 2450 | hCoV-19/England/QEUH-B2B3CF/2020\|EPI_ISL_646530\|20 | ···················· | ···················· | ···················· | ···················· | ···················· | ···················· |
| R | 2451 | hCoV-19/England/QEUH-B2B5D8/2020\|EPI_ISL_646531\|20 | ···················· | ···················· | ···················· | ···················· | ···················· | ···················· |
| R | 2452 | hCoV-19/England/ALDP-B25055/2020\|EPI_ISL_646532\|20 | ···················· | ···················· | ···················· | ···················· | ···················· | ···················· |
| R | 2453 | hCoV-19/England/ALDP-B2643A/2020\|EPI_ISL_646533\|20 | ···················· | ···················· | ···················· | ···················· | ···················· | ···················· |
| R | 2454 | hCoV-19/England/ALDP-B26230/2020\|EPI_ISL_646535\|20 | ···················· | ···················· | ···················· | ···················· | ···················· | ···················· |
| R | 2455 | hCoV-19/England/QEUH-B2C8ED/2020\|EPI_ISL_646536\|20 | ···················· | ···················· | ···················· | ···················· | ···················· | ···················· |
| R | 2456 | hCoV-19/England/ALDP-B24FB3/2020\|EPI_ISL_646537\|20 | ···················· | ·A·················· | ···················· | ···················· | ···················· | ··A················· |
| R | 2457 | hCoV-19/England/ALDP-B24EC5/2020\|EPI_ISL_646538\|20 | ···················· | ·A·················· | ···················· | ···················· | ···················· | ···················· |
| R | 2458 | hCoV-19/England/QEUH-B2BA06/2020\|EPI_ISL_646539\|20 | ···················· | ···················· | ···················· | ···················· | ···················· | ···················· |
| R | 2459 | hCoV-19/England/ALDP-B25F1C/2020\|EPI_ISL_646540\|20 | ···················· | ···················· | ···················· | ···················· | ···················· | ···················· |
| R | 2460 | hCoV-19/England/ALDP-B25107/2020\|EPI_ISL_646541\|20 | ···················· | ···················· | ···················· | ···················· | ···················· | ···················· |
| R | 2461 | hCoV-19/England/ALDP-B25F49/2020\|EPI_ISL_646542\|20 | ···················· | ···················· | ···················· | ···················· | ···················· | ···················· |
| R | 2462 | hCoV-19/England/ALDP-B25091/2020\|EPI_ISL_646543\|20 | ···················· | ···················· | ···················· | ···················· | ···················· | ···················· |
| R | 2463 | hCoV-19/England/ALDP-B252D7/2020\|EPI_ISL_646544\|20 | ···················· | ···················· | ···················· | ···················· | ···················· | ···················· |
| R | 2464 | hCoV-19/England/ALDP-B261D9/2020\|EPI_ISL_646545\|20 | ···················· | ···················· | ···················· | ···················· | ···················· | ···················· |
| R | 2465 | hCoV-19/England/ALDP-B2528C/2020\|EPI_ISL_646546\|20 | ···················· | ···················· | ···················· | ···················· | ···················· | ···················· |
| R | 2466 | hCoV-19/England/QEUH-B2BA33/2020\|EPI_ISL_646547\|20 | ···················· | ···················· | ···················· | ···················· | ···················· | ···················· |
| R | 2467 | hCoV-19/England/ALDP-B26090/2020\|EPI_ISL_646548\|20 | ···················· | ···················· | ···················· | ···················· | ···················· | ···················· |
| R | 2468 | hCoV-19/England/ALDP-B25037/2020\|EPI_ISL_646549\|20 | ···················· | ···················· | ···················· | ···················· | ···················· | ···················· |
| R | 2469 | hCoV-19/England/QEUH-B2C8FC/2020\|EPI_ISL_646550\|20 | ···················· | ···················· | ···················· | ···················· | ···················· | ···················· |
| R | 2470 | hCoV-19/England/ALDP-B26221/2020\|EPI_ISL_646551\|20 | ···················· | ···················· | ···················· | ···················· | ···················· | ···················· |
| R | 2471 | hCoV-19/England/QEUH-B2B8C0/2020\|EPI_ISL_646553\|20 | ···················· | ···················· | ···················· | ···················· | ···················· | ···················· |
| R | 2472 | hCoV-19/England/QEUH-B2BA7F/2020\|EPI_ISL_646554\|20 | ···················· | ···················· | ···················· | ···················· | ···················· | ···················· |
| R | 2473 | hCoV-19/England/ALDP-B25134/2020\|EPI_ISL_646555\|20 | ···················· | ·A·················· | ···················· | ···················· | ···················· | ···················· |
| R | 2474 | hCoV-19/England/QEUH-B2B70F/2020\|EPI_ISL_646556\|20 | ···················· | ···················· | ···················· | ···················· | ···················· | ···················· |
| R | 2475 | hCoV-19/England/ALDP-B260AF/2020\|EPI_ISL_646557\|20 | ···················· | ···················· | ···················· | ···················· | ···················· | ···················· |
| R | 2476 | hCoV-19/England/ALDP-B260EB/2020\|EPI_ISL_646558\|20 | ···················· | ···················· | ···················· | ···················· | ···················· | ···················· |
| R | 2477 | hCoV-19/England/ALDP-B24FFF/2020\|EPI_ISL_646559\|20 | ···················· | ···················· | ···················· | ···················· | ···················· | ···················· |
| R | 2478 | hCoV-19/England/ALDP-B24DC8/2020\|EPI_ISL_646561\|20 | ···················· | ···················· | ···················· | ···················· | ···················· | ···················· |
| R | 2479 | hCoV-19/England/QEUH-B2B699/2020\|EPI_ISL_646562\|20 | ···················· | ···················· | ···················· | ···················· | ···················· | ···················· |
| R | 2480 | hCoV-19/England/ALDP-B25F0D/2020\|EPI_ISL_646563\|20 | ···················· | ···················· | ···················· | ···················· | ···················· | ···················· |
| R | 2481 | hCoV-19/England/ALDP-B26203/2020\|EPI_ISL_646564\|20 | ···················· | ···················· | ···················· | ···················· | ···················· | ···················· |
| R | 2482 | hCoV-19/England/QEUH-B2B778/2020\|EPI_ISL_646565\|20 | ···················· | ···················· | ···················· | ···················· | ···················· | ···················· |
| R | 2483 | hCoV-19/England/ALDP-B25FC1/2020\|EPI_ISL_646566\|20 | ···················· | ·········T·········· | ···················· | ···················· | ···················· | ···················· |
| R | 2484 | hCoV-19/England/ALDP-B25FB2/2020\|EPI_ISL_646567\|20 | ···················· | ···················· | ···················· | ···················· | ···················· | ···················· |
| R | 2485 | hCoV-19/England/ALDP-B262E5/2020\|EPI_ISL_646568\|20 | ···················· | ·A·················· | ···················· | ···················· | ···················· | ···················· |
| R | 2486 | hCoV-19/England/ALDP-B26142/2020\|EPI_ISL_646569\|20 | ···················· | ···················· | ···················· | ···················· | ···················· | ···················· |
| R | 2487 | hCoV-19/England/QEUH-B2B4F9/2020\|EPI_ISL_646570\|20 | ···················· | ···················· | ···················· | ···················· | ···················· | ···················· |
| R | 2488 | hCoV-19/England/ALDP-B26467/2020\|EPI_ISL_646571\|20 | ···················· | ···················· | ···················· | ···················· | ···················· | ···················· |
| R | 2489 | hCoV-19/England/ALDP-B26072/2020\|EPI_ISL_646572\|20 | ···················· | ···················· | ···················· | ···················· | ···················· | ···················· |
| R | 2490 | hCoV-19/England/ALDP-B24F86/2020\|EPI_ISL_646573\|20 | ···················· | ·A·················· | ···················· | ···················· | ···················· | ··A················· |
| R | 2491 | hCoV-19/England/QEUH-B2BA24/2020\|EPI_ISL_646574\|20 | ···················· | ···················· | ···················· | ···················· | ···················· | ···················· |
| R | 2492 | hCoV-19/England/QEUH-B2B277/2020\|EPI_ISL_646575\|20 | ···················· | ···················· | ···················· | ···················· | ···················· | ···················· |
| R | 2493 | hCoV-19/England/ALDP-B264C1/2020\|EPI_ISL_646576\|20 | ···················· | ···················· | ···················· | ···················· | ···················· | ···················· |
| R | 2494 | hCoV-19/England/ALDP-B2628B/2020\|EPI_ISL_646577\|20 | ···················· | ···················· | ···················· | ···················· | ···················· | ···················· |
| R | 2495 | hCoV-19/England/QEUH-B2B620/2020\|EPI_ISL_646578\|20 | ···················· | ···················· | ···················· | ···················· | ···················· | ···················· |
| R | 2496 | hCoV-19/England/QEUH-B2CABA/2020\|EPI_ISL_646579\|20 | ···················· | ···················· | ···················· | ···················· | ···················· | ···················· |
| R | 2497 | hCoV-19/Wales/MILK-B025D7/2020\|EPI_ISL_647826\|2020 | ···················· | ···················· | ···················· | ···················· | ···················· | ···················· |
| R | 2498 | hCoV-19/Wales/MILK-B06218/2020\|EPI_ISL_647827\|2020 | ···················· | ···················· | ···················· | ···················· | ···················· | ···················· |
| R | 2499 | hCoV-19/Wales/MILK-B06306/2020\|EPI_ISL_647828\|2020 | ···················· | ···················· | ···················· | ···················· | ···················· | ···················· |
| R | 2500 | hCoV-19/Wales/MILK-B01D1C/2020\|EPI_ISL_647829\|2020 | ···················· | ···················· | ···················· | ···················· | ···················· | ···················· |
| R | 2501 | hCoV-19/Wales/MILK-B02B8A/2020\|EPI_ISL_647830\|2020 | ···················· | ···················· | ···················· | ···················· | ···················· | ···················· |
| R | 2502 | hCoV-19/Wales/MILK-B02CB4/2020\|EPI_ISL_647831\|2020 | ···················· | ···················· | ···················· | ···················· | ···················· | ···················· |
| R | 2503 | hCoV-19/Wales/MILK-B06227/2020\|EPI_ISL_647832\|2020 | ···················· | ···················· | ···················· | ···················· | ···················· | ···················· |
| R | 2504 | hCoV-19/Wales/MILK-B06360/2020\|EPI_ISL_647833\|2020 | ···················· | ···················· | ···················· | ···················· | ···················· | ···················· |
| R | 2505 | hCoV-19/Wales/MILK-B02B11/2020\|EPI_ISL_647834\|2020 | ···················· | ···················· | ···················· | ···················· | ···················· | ···················· |
| R | 2506 | hCoV-19/Wales/MILK-B02CF0/2020\|EPI_ISL_647835\|2020 | ···················· | ···················· | ···················· | ···················· | ···················· | ···················· |
| R | 2507 | hCoV-19/Wales/MILK-B0249E/2020\|EPI_ISL_647836\|2020 | ···················· | ···················· | ···················· | ···················· | ···················· | ···················· |

R 2508 hCoV-19/Wales/MILK-B02513/2020|EPI_ISL_647837|2020 .................... .................... .................... .................... .................... ....................
R 2509 hCoV-19/Wales/MILK-B02B4E/2020|EPI_ISL_647838|2020 .................... .................... .................... .................... .................... ....................
R 2510 hCoV-19/Wales/MILK-B062DC/2020|EPI_ISL_647839|2020 .................... .................... .................... .................... .................... ....................
R 2511 hCoV-19/Wales/MILK-B02BE4/2020|EPI_ISL_647840|2020 .................... .................... .................... .................... .................... ....................
R 2512 hCoV-19/Wales/MILK-B02C3C/2020|EPI_ISL_647841|2020 .................... .................... .................... .................... .................... ....................
R 2513 hCoV-19/Wales/MILK-B02C78/2020|EPI_ISL_647843|2020 .................... .................... .................... .................... .................... ....................
R 2514 hCoV-19/Wales/MILK-B01DA3/2020|EPI_ISL_647844|2020 .................... .................... .................... .................... .................... ....................
R 2515 hCoV-19/Wales/MILK-B01D2B/2020|EPI_ISL_647845|2020 .................... .................... .................... .................... .................... ....................
R 2516 hCoV-19/Wales/MILK-B06245/2020|EPI_ISL_647846|2020 .................... .................... .................... .................... .................... ....................
R 2517 hCoV-19/Wales/MILK-B02C4B/2020|EPI_ISL_647849|2020 .................... .................... .................... .................... .................... ....................
R 2518 hCoV-19/Wales/MILK-B02BC6/2020|EPI_ISL_647850|2020 .................... .................... .................... .................... .................... ....................
R 2519 hCoV-19/Wales/MILK-B02470/2020|EPI_ISL_647851|2020 .................... .................... .................... .................... .................... ....................
R 2520 hCoV-19/Wales/MILK-B02CC3/2020|EPI_ISL_647852|2020 .................... .................... .................... .................... .................... ....................
R 2521 hCoV-19/Wales/MILK-B025AA/2020|EPI_ISL_647853|2020 .................... .................... .................... .................... .................... ....................
R 2522 hCoV-19/Wales/QEUH-A8B9AE/2020|EPI_ISL_647854|2020 .................... .................... .................... .................... .................... ....................
R 2523 hCoV-19/Wales/QEUH-A8BAE7/2020|EPI_ISL_647856|2020 .................... .................... .................... .................... .................... ....................
R 2524 hCoV-19/Wales/QEUH-A8BAD8/2020|EPI_ISL_647857|2020 .................... .................... .................... .................... .................... ....................
R 2525 hCoV-19/Wales/QEUH-A8B294/2020|EPI_ISL_647858|2020 .................... .................... .................... .................... .................... ....................
R 2526 hCoV-19/Wales/QEUH-A8B59B/2020|EPI_ISL_647859|2020 .................... .................... .................... .................... .................... ....................
R 2527 hCoV-19/Wales/QEUH-A8B777/2020|EPI_ISL_647860|2020 .................... .................... .................... .................... .................... ....................
R 2528 hCoV-19/Wales/QEUH-B13175/2020|EPI_ISL_647861|2020 .................... .................... .................... .................... .................... ....................
R 2529 hCoV-19/Wales/QEUH-B121C1/2020|EPI_ISL_647862|2020 .................... .................... .................... .................... .................... ....................
R 2530 hCoV-19/Wales/QEUH-B131A2/2020|EPI_ISL_647864|2020 .................... .................... .................... .................... .................... ....................
R 2531 hCoV-19/Wales/QEUH-B131FD/2020|EPI_ISL_647866|2020 .................... .................... .................... .................... .................... ....................
R 2532 hCoV-19/Wales/QEUH-B1255C/2020|EPI_ISL_647867|2020 .................... .................... .................... .................... .................... ....................
R 2533 hCoV-19/Wales/QEUH-B1256B/2020|EPI_ISL_647868|2020 .................... .................... .................... .................... .................... ....................
R 2534 hCoV-19/Wales/QEUH-B1253E/2020|EPI_ISL_647869|2020 .................... .................... .................... .................... .................... ....................
R 2535 hCoV-19/Wales/QEUH-B1246E/2020|EPI_ISL_647871|2020 .................... .................... .................... .................... .................... ....................
R 2536 hCoV-19/Wales/QEUH-B12404/2020|EPI_ISL_647872|2020 .................... .................... .................... .................... .................... ....................
R 2537 hCoV-19/Wales/QEUH-AE2495/2020|EPI_ISL_647873|2020 .................... .................... .................... .................... .................... ....................
R 2538 hCoV-19/Wales/QEUH-AE1D13/2020|EPI_ISL_647874|2020 .................... .................... .................... .................... .................... ....................
R 2539 hCoV-19/Wales/QEUH-AE1CF8/2020|EPI_ISL_647875|2020 .................... .................... .................... .................... .................... ....................
R 2540 hCoV-19/Wales/QEUH-AE2FEF/2020|EPI_ISL_647877|2020 .................... .................... .................... .................... .................... ....................
R 2541 hCoV-19/Wales/QEUH-AE21F8/2020|EPI_ISL_647878|2020 .................... .................... .................... .................... .................... ....................
R 2542 hCoV-19/Wales/QEUH-AE23A7/2020|EPI_ISL_647879|2020 .................... .................... .................... .................... .................... ....................
R 2543 hCoV-19/Wales/QEUH-AE20CE/2020|EPI_ISL_647880|2020 .................... .................... .................... .................... .................... ....................
R 2544 hCoV-19/Wales/QEUH-AE1C34/2020|EPI_ISL_647881|2020 .................... .................... .................... .................... .................... ....................
R 2545 hCoV-19/Wales/QEUH-AE2529/2020|EPI_ISL_647882|2020 .................... .................... .................... .................... .................... ....................
R 2546 hCoV-19/Wales/QEUH-AE2E1F/2020|EPI_ISL_647883|2020 .................... .................... .................... .................... .................... ....................
R 2547 hCoV-19/Wales/QEUH-AE25CF/2020|EPI_ISL_647884|2020 .................... .................... .................... .................... .................... ....................
R 2548 hCoV-19/Wales/QEUH-AE2CAC/2020|EPI_ISL_647885|2020 .................... .................... .................... .................... .................... ....................
R 2549 hCoV-19/Wales/QEUH-AE1D22/2020|EPI_ISL_647886|2020 .................... .................... .................... .................... .................... ....................
R 2550 hCoV-19/Wales/QEUH-AE2592/2020|EPI_ISL_647887|2020 .................... .................... .................... .................... .................... ....................
R 2551 hCoV-19/Wales/QEUH-AE244A/2020|EPI_ISL_647888|2020 .................... .................... .................... .................... .................... ....................
R 2552 hCoV-19/Wales/QEUH-AE2091/2020|EPI_ISL_647889|2020 .................... .................... .................... .................... .................... ....................
R 2553 hCoV-19/Wales/QEUH-AE2019/2020|EPI_ISL_647891|2020 .................... .................... .................... .................... .................... ....................
R 2554 hCoV-19/Wales/QEUH-AE2231/2020|EPI_ISL_647892|2020 .................... .................... .................... .................... .................... ....................
R 2555 hCoV-19/Wales/QEUH-AE234D/2020|EPI_ISL_647894|2020 .................... .................... .................... .................... .................... ....................
R 2556 hCoV-19/Wales/QEUH-AE1BA0/2020|EPI_ISL_647895|2020 .................... .................... .................... .................... .................... ....................
R 2557 hCoV-19/Wales/QEUH-AE1BDD/2020|EPI_ISL_647896|2020 .................... .................... .................... .................... .................... ....................
R 2558 hCoV-19/Wales/QEUH-AE2608/2020|EPI_ISL_647897|2020 .................... .................... .................... .................... .................... ....................
R 2559 hCoV-19/Wales/QEUH-AE219E/2020|EPI_ISL_647898|2020 .................... .................... .................... .................... .................... ....................
R 2560 hCoV-19/Wales/QEUH-AE4EB3/2020|EPI_ISL_647899|2020 .................... .................... .................... .................... .................... ....................
R 2561 hCoV-19/Germany/BW-MVZ-LS-40/2020|EPI_ISL_640257|2 .................... .................... .................... .................... .................... ....................
R 2562 hCoV-19/Germany/BW-MVZ-LS-41/2020|EPI_ISL_640258|2 .................... .................... .................... .................... .................... ....................
R 2563 hCoV-19/Germany/BW-MVZ-LS-42/2020|EPI_ISL_640259|2 .................... .................... .................... .................... .................... ....................
R 2564 hCoV-19/Germany/BW-MVZ-LS-43/2020|EPI_ISL_640260|2 .................... .................... .................... .................... .................... ....................
R 2565 hCoV-19/Germany/BW-MVZ-LS-44/2020|EPI_ISL_640261|2 .................... .................... .................... .................... .................... ....................
R 2566 hCoV-19/Germany/BW-MVZ-LS-45/2020|EPI_ISL_640262|2 .................... .................... .................... .................... .................... ....................
R 2567 hCoV-19/Germany/BW-MVZ-LS-46/2020|EPI_ISL_640263|2 .................... .................... .................... .................... .................... ....................
R 2568 hCoV-19/Germany/BW-MVZ-LS-47/2020|EPI_ISL_640264|2 .................... .................... .................... .................... .................... .....G..............
R 2569 hCoV-19/USA/UT-UPHL-201123485/2020|EPI_ISL_640279| .................... .................... .................... .................... .................... ....................
R 2570 hCoV-19/USA/UT-UPHL-201117781/2020|EPI_ISL_640288| .................... .................... .................... .................... .................... ....................
R 2571 hCoV-19/USA/UT-UPHL-201128977/2020|EPI_ISL_640290| .................... .................... .................... .................... .................... ....................
R 2572 hCoV-19/USA/UT-UPHL-201118084/2020|EPI_ISL_640300| .................... .................... .................... .................... .................... ....................
R 2573 hCoV-19/Australia/VIC12844/2020|EPI_ISL_640303|202 .................... .................... .................... .................... .................... ....................
R 2575 hCoV-19/Australia/VIC16469/2020|EPI_ISL_640305|202 .................... .................... .................... .................... .................... ....................
R 2576 hCoV-19/Australia/VIC12713/2020|EPI_ISL_640308|202 .................... .................... .................... .................... .................... ....................
R 2577 hCoV-19/Australia/VIC16012/2020|EPI_ISL_640310|202 .................... .................... .................... .................... .................... ....................
R 2579 hCoV-19/Australia/VIC11990/2020|EPI_ISL_640313|202 .................... .................... .................... .................... .................... ....................
R 2580 hCoV-19/Australia/VIC12362/2020|EPI_ISL_640314|202 .................... .................... .................... .................... .................... ....................
R 2581 hCoV-19/Australia/VIC12562/2020|EPI_ISL_640315|202 .................... .................... .................... .................... .................... ....................
R 2582 hCoV-19/Australia/VIC13095/2020|EPI_ISL_640316|202 .................... .................... .................... .................... .................... ....................
R 2584 hCoV-19/Australia/VIC12171/2020|EPI_ISL_640318|202 .................... .................... .................... .................... .................... ....................
R 2586 hCoV-19/Australia/VIC16646/2020|EPI_ISL_640321|202 .................... .................... .................... .................... .................... ....................
R 2587 hCoV-19/Australia/VIC12741/2020|EPI_ISL_640322|202 .................... .................... .................... .................... .................... ....................
R 2588 hCoV-19/Australia/VIC12365/2020|EPI_ISL_640324|202 .................... .................... .................... .................... .................... ....................
R 2589 hCoV-19/Australia/VIC16175/2020|EPI_ISL_640325|202 .................... .................... .................... .................... .................... ....................
R 2590 hCoV-19/Australia/VIC12882/2020|EPI_ISL_640327|202 .................... .................... .................... .................... .................... ....................
R 2591 hCoV-19/Australia/VIC12167/2020|EPI_ISL_640328|202 .................... .................... .................... .................... .................... ....................

| | | | | | | | | |
|---|---|---|---|---|---|---|---|---|
| R | 2592 | hCoV-19/Australia/VIC12504/2020\|EPI_ISL_640329\|202 | ···················· | ···················· | ···················· | ···················· | ···················· | ···················· |
| R | 2593 | hCoV-19/Australia/VIC12789/2020\|EPI_ISL_640331\|202 | ···················· | ···················· | ···················· | ···················· | ···················· | ···················· |
| R | 2594 | hCoV-19/Australia/VIC12728/2020\|EPI_ISL_640333\|202 | ···················· | ···················· | ···················· | ···················· | ···················· | ···················· |
| R | 2595 | hCoV-19/Australia/VIC16240/2020\|EPI_ISL_640334\|202 | ···················· | ···················· | ···················· | ···················· | ···················· | ···················· |
| R | 2596 | hCoV-19/Australia/VIC12521/2020\|EPI_ISL_640335\|202 | ···················· | ···················· | ···················· | ···················· | ···················· | ···················· |
| R | 2597 | hCoV-19/Australia/VIC12642/2020\|EPI_ISL_640336\|202 | ···················· | ···················· | ···················· | ···················· | ···················· | ···················· |
| R | 2598 | hCoV-19/Australia/VIC11701/2020\|EPI_ISL_640338\|202 | ···················· | ···················· | ···················· | ···················· | ···················· | ···················· |
| R | 2599 | hCoV-19/Australia/VIC16497/2020\|EPI_ISL_640339\|202 | ···················· | ···················· | ···················· | ···················· | ···················· | ···················· |
| R | 2601 | hCoV-19/Australia/VIC16374/2020\|EPI_ISL_640341\|202 | ···················· | ···················· | ···················· | ···················· | ···················· | ···················· |
| R | 2602 | hCoV-19/Australia/VIC11723/2020\|EPI_ISL_640344\|202 | ···················· | ···················· | ···················· | ···················· | ···················· | ···················· |
| R | 2603 | hCoV-19/Australia/VIC16041/2020\|EPI_ISL_640345\|202 | ···················· | ···················· | ···················· | ···················· | ···················· | ···················· |
| R | 2604 | hCoV-19/Australia/VIC11661/2020\|EPI_ISL_640347\|202 | ···················· | ···················· | ···················· | ···················· | ···················· | ···················· |
| R | 2605 | hCoV-19/Australia/VIC12830/2020\|EPI_ISL_640348\|202 | ···················· | ···················· | ···················· | ···················· | ···················· | ···················· |
| R | 2606 | hCoV-19/Australia/VIC12495/2020\|EPI_ISL_640351\|202 | ···················· | ···················· | ···················· | ···················· | ···················· | ···················· |
| R | 2609 | hCoV-19/Australia/VIC12501/2020\|EPI_ISL_640354\|202 | ···················· | ···················· | ···················· | ···················· | ···················· | ···················· |
| R | 2610 | hCoV-19/Australia/VIC16541/2020\|EPI_ISL_640355\|202 | ···················· | ···················· | ···················· | ···················· | ···················· | ···················· |
| R | 2611 | hCoV-19/Belgium/ULG-10746/2020\|EPI_ISL_641588\|2020 | ···················· | ···················· | ···················· | ···················· | ···················· | ···················· |
| R | 2612 | hCoV-19/Belgium/ULG-10747/2020\|EPI_ISL_641589\|2020 | ···················· | ···················· | ···················· | ···················· | ···················· | ···················· |
| R | 2614 | hCoV-19/Belgium/ULG-10749/2020\|EPI_ISL_641591\|2020 | ···················· | ···················· | ···················· | ···················· | ···················· | ···················· |
| R | 2615 | hCoV-19/Belgium/ULG-10750/2020\|EPI_ISL_641592\|2020 | ···················· | ···················· | ···················· | ···················· | ···················· | ···················· |
| R | 2616 | hCoV-19/Belgium/ULG-10751/2020\|EPI_ISL_641593\|2020 | ···················· | ·A·················· | ···················· | ···················· | ···················· | ···················· |
| R | 2620 | hCoV-19/Belgium/ULG-10756/2020\|EPI_ISL_641598\|2020 | ···················· | ···················· | ···················· | ···················· | ···················· | ···················· |
| R | 2621 | hCoV-19/Belgium/ULG-10758/2020\|EPI_ISL_641600\|2020 | ···················· | ···················· | ···················· | ···················· | ···················· | ···················· |
| R | 2624 | hCoV-19/Belgium/ULG-10761/2020\|EPI_ISL_641603\|2020 | ···················· | ···················· | ···················· | ···················· | ···················· | ···················· |
| R | 2627 | hCoV-19/England/QEUH-B0F73E/2020\|EPI_ISL_641608\|20 | ···················· | ···················· | ···················· | ···················· | ···················· | ···················· |
| R | 2628 | hCoV-19/England/QEUH-B13B95/2020\|EPI_ISL_641609\|20 | ···················· | ···················· | ···················· | ···················· | ···················· | ···················· |
| R | 2629 | hCoV-19/England/QEUH-B13D9F/2020\|EPI_ISL_641610\|20 | ···················· | ···················· | ···················· | ···················· | ···················· | ···················· |
| R | 2630 | hCoV-19/England/QEUH-B13B2C/2020\|EPI_ISL_641611\|20 | ···················· | ···················· | ···················· | ···················· | ···················· | ···················· |
| R | 2631 | hCoV-19/England/QEUH-B13CDE/2020\|EPI_ISL_641613\|20 | ···················· | ···················· | ···················· | ···················· | ···················· | ···················· |
| R | 2632 | hCoV-19/England/QEUH-B13E41/2020\|EPI_ISL_641614\|20 | ···················· | ···················· | ···················· | ···················· | ···················· | ···················· |
| R | 2633 | hCoV-19/England/QEUH-B133CA/2020\|EPI_ISL_641615\|20 | ···················· | ···················· | ···················· | ···················· | ···················· | ···················· |
| R | 2634 | hCoV-19/England/QEUH-B10882/2020\|EPI_ISL_641616\|20 | ···················· | ···················· | ···················· | ···················· | ···················· | ···················· |
| R | 2635 | hCoV-19/England/QEUH-B140E0/2020\|EPI_ISL_641617\|20 | ···················· | ···················· | ···················· | ···················· | ···················· | ···················· |
| R | 2636 | hCoV-19/England/QEUH-B13FD5/2020\|EPI_ISL_641618\|20 | ···················· | ···················· | ···················· | ···················· | ···················· | ···················· |
| R | 2637 | hCoV-19/England/QEUH-B13421/2020\|EPI_ISL_641619\|20 | ···················· | ···················· | ···················· | ···················· | ···················· | ···················· |
| R | 2638 | hCoV-19/England/QEUH-B14138/2020\|EPI_ISL_641620\|20 | ···················· | ···················· | ···················· | ···················· | ···················· | ···················· |
| R | 2639 | hCoV-19/England/QEUH-B13728/2020\|EPI_ISL_641621\|20 | ···················· | ···················· | ···················· | ···················· | ···················· | ···················· |
| R | 2640 | hCoV-19/England/QEUH-B13B68/2020\|EPI_ISL_641622\|20 | ···················· | ···················· | ···················· | ···················· | ···················· | ···················· |
| R | 2641 | hCoV-19/England/QEUH-B13D26/2020\|EPI_ISL_641623\|20 | ···················· | ···················· | ···················· | ···················· | ···················· | ···················· |
| R | 2642 | hCoV-19/England/QEUH-B137DD/2020\|EPI_ISL_641625\|20 | ···················· | ···················· | ···················· | ···················· | ···················· | ···················· |
| R | 2643 | hCoV-19/England/QEUH-B1070D/2020\|EPI_ISL_641626\|20 | ···················· | ···················· | ···················· | ···················· | ···················· | ···················· |
| R | 2644 | hCoV-19/England/QEUH-B13EC9/2020\|EPI_ISL_641627\|20 | ···················· | ···················· | ···················· | ···················· | ···················· | ···················· |
| R | 2645 | hCoV-19/England/QEUH-B12C84/2020\|EPI_ISL_641629\|20 | ···················· | ···················· | ···················· | ···················· | ···················· | ···················· |
| R | 2646 | hCoV-19/England/QEUH-B10864/2020\|EPI_ISL_641630\|20 | ···················· | ···················· | ···················· | ···················· | ···················· | ···················· |
| R | 2647 | hCoV-19/England/QEUH-B134B8/2020\|EPI_ISL_641631\|20 | ···················· | ···················· | ···················· | ···················· | ···················· | ···················· |
| R | 2648 | hCoV-19/England/QEUH-B14174/2020\|EPI_ISL_641632\|20 | ···················· | ···················· | ···················· | ···················· | ···················· | ···················· |
| R | 2649 | hCoV-19/England/QEUH-B1370A/2020\|EPI_ISL_641633\|20 | ···················· | ···················· | ···················· | ···················· | ···················· | ···················· |
| R | 2650 | hCoV-19/England/QEUH-B137FB/2020\|EPI_ISL_641634\|20 | ···················· | ···················· | ···················· | ···················· | ···················· | ···················· |
| R | 2651 | hCoV-19/England/QEUH-B12D72/2020\|EPI_ISL_641635\|20 | ···················· | ···················· | ···················· | ···················· | ···················· | ···················· |
| R | 2652 | hCoV-19/England/QEUH-B14596/2020\|EPI_ISL_641636\|20 | ···················· | ···················· | ···················· | ···················· | ···················· | ···················· |
| R | 2653 | hCoV-19/England/QEUH-B13C74/2020\|EPI_ISL_641637\|20 | ···················· | ···················· | ···················· | ···················· | ···················· | ···················· |
| R | 2654 | hCoV-19/England/QEUH-B12D54/2020\|EPI_ISL_641638\|20 | ···················· | ···················· | ···················· | ···················· | ···················· | ···················· |
| R | 2655 | hCoV-19/England/QEUH-B10891/2020\|EPI_ISL_641639\|20 | ···················· | ·A·················· | ···················· | ···················· | ···················· | ···················· |
| R | 2656 | hCoV-19/England/QEUH-B107D0/2020\|EPI_ISL_641641\|20 | ···················· | ···················· | ···················· | ···················· | ···················· | ···················· |
| R | 2657 | hCoV-19/England/QEUH-B142F9/2020\|EPI_ISL_641642\|20 | ···················· | ···················· | ···················· | ···················· | ···················· | ···················· |
| R | 2658 | hCoV-19/England/QEUH-B10688/2020\|EPI_ISL_641643\|20 | ···················· | ···················· | ···················· | ···················· | ···················· | ···················· |
| R | 2659 | hCoV-19/England/QEUH-B13D71/2020\|EPI_ISL_641644\|20 | ···················· | ···················· | ···················· | ···················· | ···················· | ···················· |
| R | 2660 | hCoV-19/England/QEUH-B14280/2020\|EPI_ISL_641645\|20 | ···················· | ···················· | ···················· | ···················· | ···················· | ···················· |
| R | 2661 | hCoV-19/England/QEUH-B1353D/2020\|EPI_ISL_641646\|20 | ···················· | ···················· | ···················· | ···················· | ···················· | ···················· |
| R | 2662 | hCoV-19/England/QEUH-B0F0AC/2020\|EPI_ISL_641647\|20 | ···················· | ···················· | ···················· | ···················· | ···················· | ···················· |
| R | 2663 | hCoV-19/England/QEUH-B0F006/2020\|EPI_ISL_641648\|20 | ···················· | ···················· | ···················· | ···················· | ···················· | ···················· |
| R | 2664 | hCoV-19/England/QEUH-B13B59/2020\|EPI_ISL_641649\|20 | ···················· | ···················· | ···················· | ···················· | ···················· | ···················· |
| R | 2665 | hCoV-19/England/QEUH-B13861/2020\|EPI_ISL_641650\|20 | ···················· | ···················· | ···················· | ···················· | ···················· | ···················· |
| R | 2666 | hCoV-19/England/QEUH-B0F0BB/2020\|EPI_ISL_641651\|20 | ···················· | ···················· | ···················· | ···················· | ···················· | ···················· |
| R | 2667 | hCoV-19/England/QEUH-B1400E/2020\|EPI_ISL_641652\|20 | ···················· | ···················· | ···················· | ···················· | ···················· | ···················· |
| R | 2668 | hCoV-19/England/QEUH-B10855/2020\|EPI_ISL_641653\|20 | ···················· | ···················· | ···················· | ···················· | ···················· | ···················· |
| R | 2669 | hCoV-19/England/QEUH-B0F7D4/2020\|EPI_ISL_641655\|20 | ···················· | ···················· | ···················· | ···················· | ···················· | ···················· |
| R | 2670 | hCoV-19/England/QEUH-B12D27/2020\|EPI_ISL_641656\|20 | ···················· | ···················· | ···················· | ···················· | ···················· | ···················· |
| R | 2671 | hCoV-19/England/QEUH-B14147/2020\|EPI_ISL_641657\|20 | ···················· | ···················· | ···················· | ···················· | ···················· | ···················· |
| R | 2672 | hCoV-19/England/QEUH-B0F0CA/2020\|EPI_ISL_641658\|20 | ···················· | ···················· | ···················· | ···················· | ···················· | ···················· |
| R | 2673 | hCoV-19/England/QEUH-B13D08/2020\|EPI_ISL_641659\|20 | ···················· | ···················· | ···················· | ···················· | ···T················ | ···················· |
| R | 2674 | hCoV-19/England/QEUH-B0F7F2/2020\|EPI_ISL_641660\|20 | ···················· | ···················· | ···················· | ···················· | ···················· | ···················· |
| R | 2675 | hCoV-19/England/QEUH-B0F76B/2020\|EPI_ISL_641662\|20 | ···················· | ···················· | ···················· | ···················· | ···················· | ···················· |
| R | 2676 | hCoV-19/England/QEUH-B142CC/2020\|EPI_ISL_641663\|20 | ···················· | ···················· | ···················· | ···················· | ···················· | ···················· |
| R | 2677 | hCoV-19/England/QEUH-B140B3/2020\|EPI_ISL_641665\|20 | ···················· | ···················· | ···················· | ···················· | ···················· | ···················· |
| R | 2678 | hCoV-19/England/QEUH-B0F7B6/2020\|EPI_ISL_641666\|20 | ···················· | ···················· | ···················· | ···················· | ···················· | ···················· |
| R | 2679 | hCoV-19/England/QEUH-B138BC/2020\|EPI_ISL_641667\|20 | ···················· | ···················· | ···················· | ···················· | ···················· | ···················· |
| R | 2680 | hCoV-19/England/QEUH-B12CA2/2020\|EPI_ISL_641668\|20 | ···················· | ···················· | ···················· | ···················· | ···················· | ···················· |
| R | 2681 | hCoV-19/England/QEUH-B1401D/2020\|EPI_ISL_641669\|20 | ···················· | ···················· | ···················· | ···················· | ···················· | ···················· |
| R | 2682 | hCoV-19/England/QEUH-B102FD/2020\|EPI_ISL_641670\|20 | ···················· | ···················· | ···················· | ···················· | ···················· | ···················· |

| | | | | | | | |
|---|---|---|---|---|---|---|---|
| R | 2683 hCoV-19/England/QEUH-B13A01/2020\|EPI_ISL_641671\|20 | ··················· | ··················· | ···················· | ··················· | ··················· | ··················· |
| R | 2684 hCoV-19/England/QEUH-B141ED/2020\|EPI_ISL_641673\|20 | ··················· | ··················· | ···················· | ··················· | ··················· | ··················· |
| R | 2685 hCoV-19/England/QEUH-B0F798/2020\|EPI_ISL_641674\|20 | ··················· | ··················· | ···················· | ··················· | ··················· | ··················· |
| R | 2686 hCoV-19/England/QEUH-B13F3F/2020\|EPI_ISL_641675\|20 | ··················· | ··················· | ···················· | ··················· | ··················· | ··················· |
| R | 2687 hCoV-19/England/QEUH-B1360D/2020\|EPI_ISL_641676\|20 | ··················· | ··················· | ···················· | ··················· | ··················· | ··················· |
| R | 2688 hCoV-19/England/QEUH-B10846/2020\|EPI_ISL_641677\|20 | ··················· | ··················· | ···················· | ··················· | ··················· | ··················· |
| R | 2689 hCoV-19/England/QEUH-B107C1/2020\|EPI_ISL_641678\|20 | ··················· | ··················· | ···················· | ··················· | ··················· | ··················· |
| R | 2690 hCoV-19/England/QEUH-B0EFCE/2020\|EPI_ISL_641679\|20 | ··················· | ··················· | ···················· | ··················· | ··················· | ··················· |
| R | 2691 hCoV-19/England/QEUH-B14183/2020\|EPI_ISL_641680\|20 | ··················· | ··················· | ···················· | ··················· | ··················· | ··················· |
| R | 2692 hCoV-19/England/QEUH-B13CA1/2020\|EPI_ISL_641681\|20 | ··················· | ··················· | ···················· | ··················· | ·····T············· | ··················· |
| R | 2693 hCoV-19/England/QEUH-B0F82C/2020\|EPI_ISL_641682\|20 | ··················· | ··················· | ···················· | ··················· | ··················· | ··················· |
| R | 2694 hCoV-19/England/QEUH-B0EFEC/2020\|EPI_ISL_641683\|20 | ··················· | ··················· | ···················· | ··················· | ··················· | ··················· |
| R | 2695 hCoV-19/England/QEUH-B1305A/2020\|EPI_ISL_641684\|20 | ··················· | ·A················· | ···················· | ··················· | ··················· | ··················· |
| R | 2696 hCoV-19/England/QEUH-B13CFC/2020\|EPI_ISL_641685\|20 | ··················· | ··················· | ···················· | ··················· | ··················· | ··················· |
| R | 2697 hCoV-19/England/QEUH-B12F7C/2020\|EPI_ISL_641686\|20 | ··················· | ··················· | ···················· | ··················· | ··················· | ··················· |
| R | 2698 hCoV-19/England/MILK-B02A50/2020\|EPI_ISL_642930\|20 | ··················· | ··················· | ···················· | ··················· | ··················· | ··················· |
| R | 2699 hCoV-19/England/MILK-B02C87/2020\|EPI_ISL_642931\|20 | ··················· | ··················· | ···················· | ··················· | ··················· | ··················· |
| R | 2700 hCoV-19/England/MILK-B0215B/2020\|EPI_ISL_642932\|20 | ··················· | ··················· | ···················· | ··················· | ··················· | ··················· |
| R | 2701 hCoV-19/England/MILK-B028B0/2020\|EPI_ISL_642933\|20 | ··················· | ··················· | ···················· | ··················· | ··················· | ··················· |
| R | 2702 hCoV-19/England/MILK-B020A9/2020\|EPI_ISL_642934\|20 | ··················· | ··················· | ···················· | ··················· | ··················· | ··················· |
| R | 2703 hCoV-19/England/MILK-B06139/2020\|EPI_ISL_642937\|20 | ··················· | ··················· | ···················· | ··················· | ·····T············· | ··················· |
| R | 2704 hCoV-19/England/MILK-B02C69/2020\|EPI_ISL_642939\|20 | ··················· | ··················· | ···················· | ··················· | ··················· | ··················· |
| R | 2705 hCoV-19/Wales/MILK-B02768/2020\|EPI_ISL_642942\|2020 | ··················· | ··················· | ···················· | ··················· | ··················· | ··················· |
| R | 2706 hCoV-19/England/MILK-B05F4F/2020\|EPI_ISL_642945\|20 | ··················· | ··················· | ···················· | ··················· | ··················· | ··················· |
| R | 2707 hCoV-19/England/MILK-B02917/2020\|EPI_ISL_642946\|20 | ··················· | ··················· | ···················· | ··················· | ··················· | ··················· |
| R | 2708 hCoV-19/England/MILK-B01E46/2020\|EPI_ISL_642947\|20 | ··················· | ··················· | ···················· | ··················· | ··················· | ··················· |
| R | 2709 hCoV-19/England/MILK-B02197/2020\|EPI_ISL_642948\|20 | ··················· | ··················· | ···················· | ··················· | ··················· | ··················· |
| R | 2710 hCoV-19/England/MILK-B01D76/2020\|EPI_ISL_642949\|20 | ··················· | ··················· | ···················· | ··················· | ··················· | ··················· |
| R | 2711 hCoV-19/England/MILK-B02865/2020\|EPI_ISL_642950\|20 | ··················· | ··················· | ···················· | ··················· | ··················· | ··················· |
| R | 2712 hCoV-19/England/MILK-B01D85/2020\|EPI_ISL_642951\|20 | ··················· | ··················· | ···················· | ··················· | ··················· | ··················· |
| R | 2713 hCoV-19/England/MILK-B0637F/2020\|EPI_ISL_642952\|20 | ··················· | ··················· | ···················· | ··················· | ·····T············· | ··················· |
| R | 2714 hCoV-19/England/MILK-B06290/2020\|EPI_ISL_642953\|20 | ··················· | ··················· | ···················· | ··················· | ··················· | ··················· |
| R | 2715 hCoV-19/England/MILK-B02953/2020\|EPI_ISL_642954\|20 | ··················· | ··················· | ···················· | ··················· | ··················· | ··················· |
| R | 2716 hCoV-19/England/MILK-B01E64/2020\|EPI_ISL_642955\|20 | ··················· | ··················· | ···················· | ··················· | ··················· | ··················· |
| R | 2717 hCoV-19/England/MILK-B01FE9/2020\|EPI_ISL_642956\|20 | ··················· | ··················· | ···················· | ··················· | ··················· | ··················· |
| R | 2718 hCoV-19/England/MILK-B02021/2020\|EPI_ISL_642957\|20 | ··················· | ··················· | ···················· | ··················· | ··················· | ··················· |
| R | 2719 hCoV-19/England/MILK-B02285/2020\|EPI_ISL_642958\|20 | ··················· | ··················· | ···················· | ··················· | ··················· | ··················· |
| R | 2720 hCoV-19/England/MILK-B05ED9/2020\|EPI_ISL_642959\|20 | ··················· | ··················· | ···················· | ··················· | ··················· | ··················· |
| R | 2721 hCoV-19/England/MILK-B0267A/2020\|EPI_ISL_642960\|20 | ··················· | ··················· | ···················· | ··················· | ··················· | ··················· |
| R | 2722 hCoV-19/England/MILK-B0222B/2020\|EPI_ISL_642961\|20 | ··················· | ··················· | ···················· | ··················· | ··················· | ··················· |
| R | 2723 hCoV-19/England/MILK-B05EF7/2020\|EPI_ISL_642962\|20 | ··················· | ··················· | ···················· | ··················· | ··················· | ··················· |
| R | 2724 hCoV-19/England/MILK-B02ABA/2020\|EPI_ISL_642963\|20 | ··················· | ··················· | ···················· | ··················· | ··················· | ··················· |
| R | 2725 hCoV-19/England/MILK-B06096/2020\|EPI_ISL_642964\|20 | ··················· | ··················· | ···················· | ··················· | ··················· | ··················· |
| R | 2726 hCoV-19/England/MILK-B01FCB/2020\|EPI_ISL_642965\|20 | ··················· | ··················· | ···················· | ··················· | ··················· | ··················· |
| R | 2727 hCoV-19/England/MILK-B06193/2020\|EPI_ISL_642966\|20 | ··················· | ··················· | ···················· | ··················· | ··················· | ··················· |
| R | 2728 hCoV-19/England/MILK-B02759/2020\|EPI_ISL_642967\|20 | ··················· | ··················· | ···················· | ··················· | ··················· | ··················· |
| R | 2729 hCoV-19/England/MILK-B06254/2020\|EPI_ISL_642968\|20 | ··················· | ··················· | ···················· | ··················· | ··················· | ··················· |
| R | 2730 hCoV-19/England/MILK-B01EA0/2020\|EPI_ISL_642969\|20 | ··················· | ··················· | ···················· | ··················· | ··················· | ··················· |
| R | 2731 hCoV-19/England/MILK-B031B4/2020\|EPI_ISL_642970\|20 | ··················· | ··················· | ···················· | ··················· | ··················· | ··················· |
| R | 2732 hCoV-19/England/MILK-B02962/2020\|EPI_ISL_642971\|20 | ··················· | ··················· | ···················· | ··················· | ··················· | ··················· |
| R | 2733 hCoV-19/England/MILK-B0206D/2020\|EPI_ISL_642972\|20 | ··················· | ··················· | ···················· | ··················· | ··················· | ··················· |
| R | 2734 hCoV-19/England/MILK-B025F5/2020\|EPI_ISL_642973\|20 | ··················· | ··················· | ···················· | ··················· | ··················· | ··················· |
| R | 2735 hCoV-19/England/MILK-B060B4/2020\|EPI_ISL_642974\|20 | ··················· | ··················· | ···················· | ··················· | ·····T············· | ··················· |
| R | 2736 hCoV-19/England/MILK-B061FD/2020\|EPI_ISL_642975\|20 | ··················· | ··················· | ···················· | ··················· | ··················· | ··················· |
| R | 2737 hCoV-19/England/MILK-B029BD/2020\|EPI_ISL_642976\|20 | ··················· | ··················· | ···················· | ··················· | ··················· | ··················· |
| R | 2738 hCoV-19/England/MILK-B06333/2020\|EPI_ISL_642977\|20 | ··················· | ··················· | ···················· | ··················· | ··················· | ··················· |
| R | 2739 hCoV-19/England/MILK-B0611B/2020\|EPI_ISL_642978\|20 | ··················· | ··················· | ···················· | ··················· | ·····T············· | ··················· |
| R | 2740 hCoV-19/England/MILK-B061B1/2020\|EPI_ISL_642979\|20 | ··················· | ··················· | ···················· | ··················· | ··················· | ··················· |
| R | 2741 hCoV-19/England/MILK-B05E15/2020\|EPI_ISL_642980\|20 | ··················· | ··················· | ···················· | ··················· | ·····T············· | ··················· |
| R | 2742 hCoV-19/England/MILK-B021C4/2020\|EPI_ISL_642981\|20 | ··················· | ·A················· | ···················· | ··················· | ··················· | ··················· |
| R | 2743 hCoV-19/England/MILK-B0207C/2020\|EPI_ISL_642982\|20 | ··················· | ··················· | ···················· | ··················· | ··················· | ··················· |
| R | 2744 hCoV-19/England/MILK-B02391/2020\|EPI_ISL_642983\|20 | ··················· | ··················· | ···················· | ··················· | ··················· | ··················· |
| R | 2745 hCoV-19/England/MILK-B02407/2020\|EPI_ISL_642984\|20 | ··················· | ··················· | ···················· | ··················· | ··················· | ··················· |
| R | 2746 hCoV-19/England/MILK-B02935/2020\|EPI_ISL_642985\|20 | ··················· | ··················· | ···················· | ··················· | ··················· | ··················· |
| R | 2747 hCoV-19/England/MILK-B05E51/2020\|EPI_ISL_642986\|20 | ··················· | ··················· | ···················· | ··················· | ·····T············· | ··················· |
| R | 2748 hCoV-19/England/MILK-B0307B/2020\|EPI_ISL_642987\|20 | ··················· | ··················· | ···················· | ··················· | ··················· | ··················· |
| R | 2749 hCoV-19/England/MILK-B02373/2020\|EPI_ISL_642988\|20 | ··················· | ··················· | ···················· | ··················· | ··················· | ··················· |
| R | 2750 hCoV-19/England/MILK-B0601E/2020\|EPI_ISL_642989\|20 | ··················· | ··················· | ···················· | ··················· | ··················· | ··················· |
| R | 2751 hCoV-19/England/MILK-B02908/2020\|EPI_ISL_642990\|20 | ··················· | ··················· | ···················· | ··················· | ··················· | ··················· |
| R | 2752 hCoV-19/England/MILK-B02179/2020\|EPI_ISL_642991\|20 | ··················· | ··················· | ···················· | ··················· | ·····T············· | ··················· |
| R | 2753 hCoV-19/England/MILK-B05E33/2020\|EPI_ISL_642992\|20 | ··················· | ··················· | ···················· | ··················· | ··················· | ··················· |
| R | 2754 hCoV-19/England/MILK-B029DB/2020\|EPI_ISL_642993\|20 | ··················· | ··················· | ···················· | ··················· | ··················· | ··················· |
| R | 2755 hCoV-19/England/MILK-B024AD/2020\|EPI_ISL_642994\|20 | ··················· | ··················· | ···················· | ··················· | ··················· | ··················· |
| R | 2756 hCoV-19/England/MILK-B02786/2020\|EPI_ISL_642995\|20 | ··················· | ··················· | ···················· | ··················· | ··················· | ··················· |
| R | 2757 hCoV-19/England/MILK-B0214C/2020\|EPI_ISL_642996\|20 | ··················· | ··················· | ···················· | ··················· | ··················· | ··················· |
| R | 2758 hCoV-19/England/MILK-B06324/2020\|EPI_ISL_642997\|20 | ··················· | ··················· | ···················· | ··················· | ··················· | ··················· |
| R | 2759 hCoV-19/England/MILK-B0271D/2020\|EPI_ISL_642998\|20 | ··················· | ··················· | ···················· | ··················· | ··················· | ··················· |
| R | 2760 hCoV-19/England/MILK-B05FE5/2020\|EPI_ISL_642999\|20 | ··················· | ·A················· | ···················· | ··················· | ··················· | ··A················ |
| R | 2761 hCoV-19/Singapore/1403/2020\|EPI_ISL_645130\|2020-11 | ··················· | ·A················· | ···················· | ··················· | ··················· | ··A················ |
| R | 2762 hCoV-19/Singapore/1404/2020\|EPI_ISL_645131\|2020-11 | ··················· | ·A················· | ···················· | ··················· | ··················· | ··A················ |

| | | | | | | | |
|---|---|---|---|---|---|---|---|
| R | 2763 hCoV-19/Singapore/1405/2020\|EPI_ISL_645132\|2020-11 | ···················· | ···················· | ···················· | ···················· | ···················· | ···················· |
| R | 2764 hCoV-19/England/204720404/2020\|EPI_ISL_645134\|2020 | ···················· | ···················· | ···················· | ···················· | ···················· | ···················· |
| R | 2765 hCoV-19/England/204720405/2020\|EPI_ISL_645135\|2020 | ···················· | ···················· | ···················· | ···················· | ···················· | ···················· |
| R | 2766 hCoV-19/England/204720406/2020\|EPI_ISL_645136\|2020 | ···················· | ···················· | ···················· | ···················· | ···················· | ···················· |
| R | 2767 hCoV-19/France/NAQ-SC203/2020\|EPI_ISL_645169\|2020- | ···················· | ···················· | ···················· | ···················· | ···················· | ···················· |
| R | 2768 hCoV-19/England/QEUH-B2BA9D/2020\|EPI_ISL_646460\|20 | ···················· | ···················· | ···················· | ···················· | ···················· | ···················· |
| R | 2769 hCoV-19/England/QEUH-B2B8B1/2020\|EPI_ISL_646461\|20 | ···················· | ···················· | ···················· | ···················· | ···················· | ···················· |
| R | 2770 hCoV-19/England/QEUH-B2B356/2020\|EPI_ISL_646462\|20 | ···················· | ···················· | ···················· | ···················· | ···················· | ···················· |
| R | 2771 hCoV-19/England/ALDP-B26379/2020\|EPI_ISL_646463\|20 | ···················· | ···················· | ···················· | ···················· | ···················· | ···················· |
| R | 2772 hCoV-19/England/QEUH-B2B64E/2020\|EPI_ISL_646464\|20 | ···················· | ···················· | ···················· | ···················· | ···················· | ···················· |
| R | 2773 hCoV-19/England/QEUH-B2CAE7/2020\|EPI_ISL_646465\|20 | ···················· | ···················· | ···················· | ···················· | ···················· | ···················· |
| R | 2774 hCoV-19/England/ALDP-B250FB/2020\|EPI_ISL_646466\|20 | ···················· | ···················· | ···················· | ···················· | ···················· | ···················· |
| R | 2775 hCoV-19/England/ALDP-B264B2/2020\|EPI_ISL_646467\|20 | ···················· | ···················· | ···················· | ···················· | ···················· | ···················· |
| R | 2776 hCoV-19/England/ALDP-B24EE3/2020\|EPI_ISL_646468\|20 | ···················· | ···················· | ···················· | ···················· | ···················· | ···················· |
| R | 2777 hCoV-19/Scotland/QEUH-B2B5F6/2020\|EPI_ISL_646469\|2 | ···················· | ···················· | ···················· | ···················· | ···················· | ···················· |
| R | 2778 hCoV-19/England/QEUH-B2C9DB/2020\|EPI_ISL_646470\|20 | ···················· | ···················· | ···················· | ···················· | ···················· | ···················· |
| R | 2779 hCoV-19/England/QEUH-B2B259/2020\|EPI_ISL_646471\|20 | ···················· | ···················· | ···················· | ···················· | ···················· | ···················· |
| R | 2780 hCoV-19/England/QEUH-B2B2FF/2020\|EPI_ISL_646472\|20 | ···················· | ···················· | ···················· | ···················· | ···················· | ···················· |
| R | 2781 hCoV-19/England/QEUH-B2C99F/2020\|EPI_ISL_646473\|20 | ···················· | ···················· | ···················· | ···················· | ···················· | ···················· |
| R | 2782 hCoV-19/England/QEUH-B2B9CD/2020\|EPI_ISL_646474\|20 | ···················· | ···················· | ···················· | ···················· | ···················· | ···················· |
| R | 2783 hCoV-19/England/QEUH-B2B286/2020\|EPI_ISL_646475\|20 | ···················· | ···················· | ···················· | ···················· | ···················· | ···················· |
| R | 2784 hCoV-19/England/ALDP-B25073/2020\|EPI_ISL_646476\|20 | ···················· | ···················· | ···················· | ···················· | ···················· | ···················· |
| R | 2785 hCoV-19/England/ALDP-B26018/2020\|EPI_ISL_646478\|20 | ···················· | ···················· | ···················· | ···················· | ···················· | ···················· |
| R | 2786 hCoV-19/England/ALDP-B26045/2020\|EPI_ISL_646479\|20 | ···················· | ···················· | ···················· | ···················· | ···················· | ···················· |
| R | 2787 hCoV-19/England/QEUH-B2BB12/2020\|EPI_ISL_646480\|20 | ···················· | ···················· | ···················· | ···················· | ···················· | ···················· |
| R | 2788 hCoV-19/England/QEUH-B2B5E7/2020\|EPI_ISL_646481\|20 | ···················· | ···················· | ···················· | ···················· | ···················· | ···················· |
| R | 2789 hCoV-19/England/ALDP-B25143/2020\|EPI_ISL_646486\|20 | ···················· | ·A·················· | ···················· | ···················· | ···················· | ···················· |
| R | 2790 hCoV-19/England/ALDP-B24D6E/2020\|EPI_ISL_646487\|20 | ···················· | ···················· | ···················· | ···················· | ···················· | ···················· |
| R | 2791 hCoV-19/England/ALDP-B25082/2020\|EPI_ISL_646488\|20 | ···················· | ···················· | ···················· | ···················· | ···················· | ···················· |
| R | 2792 hCoV-19/England/ALDP-B252B9/2020\|EPI_ISL_646489\|20 | ···················· | ···················· | ···················· | ···················· | ···················· | ···················· |
| R | 2793 hCoV-19/England/ALDP-B2626D/2020\|EPI_ISL_646490\|20 | ···················· | ·A·················· | ···················· | ···················· | ···················· | ···················· |
| R | 2794 hCoV-19/England/ALDP-B25231/2020\|EPI_ISL_646491\|20 | ···················· | ···················· | ···················· | ···················· | ···················· | ···················· |
| R | 2795 hCoV-19/England/ALDP-B261AC/2020\|EPI_ISL_646492\|20 | ···················· | ···················· | ···················· | ···················· | ···················· | ···················· |
| R | 2796 hCoV-19/England/ALDP-B25204/2020\|EPI_ISL_646493\|20 | ···················· | ···················· | ···················· | ···················· | ···················· | ···················· |
| R | 2797 hCoV-19/England/ALDP-B262A9/2020\|EPI_ISL_646494\|20 | ···················· | ···················· | ···················· | ···················· | ···················· | ···················· |
| R | 2798 hCoV-19/England/QEUH-B2C926/2020\|EPI_ISL_646495\|20 | ···················· | ···················· | ···················· | ···················· | ···················· | ···················· |
| R | 2799 hCoV-19/England/QEUH-B2B909/2020\|EPI_ISL_646496\|20 | ···················· | ···················· | ···················· | ···················· | ···················· | ···················· |
| R | 2800 hCoV-19/England/ALDP-B2525F/2020\|EPI_ISL_646497\|20 | ···················· | ···················· | ···················· | ···················· | ···················· | ···················· |
| R | 2801 hCoV-19/England/QEUH-B2B6E4/2020\|EPI_ISL_646498\|20 | ···················· | ···················· | ···················· | ···················· | ···················· | ···················· |
| R | 2802 hCoV-19/England/QEUH-B2B4EA/2020\|EPI_ISL_646499\|20 | ···················· | ···················· | ···················· | ···················· | ···················· | ···················· |
| R | 2803 hCoV-19/England/ALDP-B1A9B2/2020\|EPI_ISL_647700\|20 | ···················· | ···················· | ···················· | ···················· | ···················· | ···················· |
| R | 2804 hCoV-19/England/CAMC-B2001E/2020\|EPI_ISL_647701\|20 | ···················· | ···················· | ···················· | ···················· | ···················· | ···················· |
| R | 2805 hCoV-19/England/CAMC-B20E41/2020\|EPI_ISL_647702\|20 | ···················· | ···················· | ···················· | ···················· | ···················· | ···················· |
| R | 2806 hCoV-19/England/CAMC-B20B2C/2020\|EPI_ISL_647703\|20 | ···················· | ·A·················· | ···················· | ···················· | ···················· | ···················· |
| R | 2807 hCoV-19/England/CAMC-B20D53/2020\|EPI_ISL_647790\|20 | ···················· | ···················· | ···················· | ···················· | ···················· | ···················· |
| R | 2808 hCoV-19/England/CAMC-B1FFF7/2020\|EPI_ISL_647791\|20 | ···················· | ···················· | ···················· | ···················· | ···················· | ···················· |
| R | 2809 hCoV-19/England/ALDP-B1A5DC/2020\|EPI_ISL_647792\|20 | ···················· | ···················· | ···················· | ···················· | ···················· | ···················· |
| R | 2810 hCoV-19/England/CAMC-B205F1/2020\|EPI_ISL_647793\|20 | ···················· | ···················· | ···················· | ···················· | ···················· | ···················· |
| R | 2811 hCoV-19/England/CAMC-B212CC/2020\|EPI_ISL_647794\|20 | ···················· | ···················· | ···················· | ···················· | ···················· | ···················· |
| R | 2812 hCoV-19/England/CAMC-B1FDFD/2020\|EPI_ISL_647795\|20 | ···················· | ···················· | ···················· | ···················· | ···················· | ···················· |
| R | 2813 hCoV-19/England/CAMC-B1FDC0/2020\|EPI_ISL_647796\|20 | ···················· | ·A·················· | ···················· | ···················· | ···················· | ···A················ |
| R | 2814 hCoV-19/Wales/QEUH-B13913/2020\|EPI_ISL_647797\|2020 | ···················· | ···················· | ···················· | ···················· | ···················· | ···················· |
| R | 2815 hCoV-19/Wales/QEUH-B138F8/2020\|EPI_ISL_647798\|2020 | ···················· | ···················· | ···················· | ···················· | ···················· | ···················· |
| R | 2816 hCoV-19/Wales/QEUH-B13834/2020\|EPI_ISL_647799\|2020 | ···················· | ···················· | ···················· | ···················· | ···················· | ···················· |
| R | 2817 hCoV-19/Germany/BW-MVZ-LS-17/2020\|EPI_ISL_640235\|2 | ···················· | ···················· | ···················· | ···················· | ···················· | ···················· |
| R | 2818 hCoV-19/Germany/BW-MVZ-LS-18/2020\|EPI_ISL_640236\|2 | ···················· | ···················· | ···················· | ···················· | ···················· | ···················· |
| R | 2819 hCoV-19/Germany/BW-MVZ-LS-20/2020\|EPI_ISL_640238\|2 | ···················· | ···················· | ···················· | ···················· | ···················· | ······G············· |
| R | 2820 hCoV-19/Germany/BW-MVZ-LS-21/2020\|EPI_ISL_640239\|2 | ···················· | ···················· | ···················· | ···················· | ···················· | ···················· |
| R | 2821 hCoV-19/Germany/BW-MVZ-LS-22/2020\|EPI_ISL_640240\|2 | ···················· | ···················· | ···················· | ···················· | ···················· | ···················· |
| R | 2822 hCoV-19/Germany/BW-MVZ-LS-23/2020\|EPI_ISL_640241\|2 | ···················· | ···················· | ···················· | ···················· | ···················· | ···················· |
| R | 2823 hCoV-19/Germany/BW-MVZ-LS-24/2020\|EPI_ISL_640242\|2 | ···················· | ···················· | ···················· | ···················· | ···················· | ···················· |
| R | 2824 hCoV-19/Germany/BW-MVZ-LS-25/2020\|EPI_ISL_640243\|2 | ···················· | ···················· | ···················· | ···················· | ···················· | ···················· |
| R | 2825 hCoV-19/Germany/BW-MVZ-LS-26/2020\|EPI_ISL_640244\|2 | ···················· | ···················· | ···················· | ···················· | ···················· | ···················· |
| R | 2826 hCoV-19/Germany/BW-MVZ-LS-27/2020\|EPI_ISL_640245\|2 | ···················· | ···················· | ···················· | ···················· | ···················· | ······G············· |
| R | 2827 hCoV-19/Germany/BW-MVZ-LS-28/2020\|EPI_ISL_640246\|2 | ···················· | ···················· | ···················· | ···················· | ···················· | ···················· |
| R | 2828 hCoV-19/Germany/BW-MVZ-LS-29/2020\|EPI_ISL_640247\|2 | ···················· | ···················· | ···················· | ···················· | ···················· | ······G············· |
| R | 2829 hCoV-19/Germany/BW-MVZ-LS-30/2020\|EPI_ISL_640248\|2 | ···················· | ···················· | ···················· | ···················· | ···················· | ···················· |
| R | 2830 hCoV-19/Germany/BW-MVZ-LS-34/2020\|EPI_ISL_640251\|2 | ···················· | ···················· | ···················· | ···················· | ···················· | ···················· |
| R | 2831 hCoV-19/Germany/BW-MVZ-LS-35/2020\|EPI_ISL_640252\|2 | ···················· | ···················· | ···················· | ···················· | ···················· | ···················· |
| R | 2832 hCoV-19/Germany/BW-MVZ-LS-36/2020\|EPI_ISL_640253\|2 | ···················· | ···················· | ···················· | ···················· | ···················· | ···················· |
| R | 2833 hCoV-19/Germany/BW-MVZ-LS-37/2020\|EPI_ISL_640254\|2 | ···················· | ···················· | ···················· | ···················· | ···················· | ···················· |
| R | 2834 hCoV-19/Germany/BW-MVZ-LS-38/2020\|EPI_ISL_640255\|2 | ···················· | ···················· | ···················· | ···················· | ···················· | ···················· |
| R | 2835 hCoV-19/Germany/BW-MVZ-LS-39/2020\|EPI_ISL_640256\|2 | ···················· | ···················· | ···················· | ···················· | ···················· | ······G············· |
| R | 2836 hCoV-19/Germany/BW-MVZ-LS-51/2020\|EPI_ISL_640268\|2 | ···················· | ···················· | ···················· | ···················· | ···················· | ···················· |
| R | 2837 hCoV-19/Germany/BW-MVZ-LS-54/2020\|EPI_ISL_640271\|2 | ···················· | ···················· | ···················· | ···················· | ···················· | ···················· |
| R | 2841 hCoV-19/Belgium/ULG-10726/2020\|EPI_ISL_641568\|2020 | ···················· | ···················· | ···················· | ···················· | ···················· | ···················· |
| R | 2850 hCoV-19/Belgium/ULG-10735/2020\|EPI_ISL_641577\|2020 | ···················· | ···················· | ···················· | ···················· | ···················· | ···················· |
| R | 2851 hCoV-19/Belgium/ULG-10736/2020\|EPI_ISL_641578\|2020 | ···················· | ···················· | ···················· | ···················· | ···················· | ···················· |
| R | 2854 hCoV-19/Belgium/ULG-10739/2020\|EPI_ISL_641581\|2020 | ···················· | ···················· | ···················· | ···················· | ···················· | ···················· |
| R | 2856 hCoV-19/Belgium/ULG-10742/2020\|EPI_ISL_641584\|2020 | ···················· | ···················· | ···················· | ···················· | ···················· | ···················· |

| | | | | | | | |
|---|---|---|---|---|---|---|---|
| R | 2858 hCoV-19/Belgium/ULG-10744/2020\|EPI_ISL_641586\|2020 | .................... | .................... | ..................... | .................... | .................... | .................... |
| R | 2859 hCoV-19/Belgium/ULG-10745/2020\|EPI_ISL_641587\|2020 | .................... | .................... | ..................... | .................... | .................... | .................... |
| R | 2861 hCoV-19/England/MILK-B060D2/2020\|EPI_ISL_642800\|20 | .................... | .................... | ..................... | .................... | .................... | .................... |
| R | 2862 hCoV-19/England/MILK-B01CE2/2020\|EPI_ISL_642801\|20 | .................... | .................... | ..................... | .................... | .................... | .................... |
| R | 2863 hCoV-19/England/MILK-B028CF/2020\|EPI_ISL_642802\|20 | .................... | .................... | ..................... | .................... | .................... | .................... |
| R | 2864 hCoV-19/England/MILK-B01D94/2020\|EPI_ISL_642803\|20 | .................... | .................... | ..................... | .................... | .................... | .................... |
| R | 2865 hCoV-19/England/MILK-B06078/2020\|EPI_ISL_642804\|20 | .................... | .................... | ..................... | .................... | .....T.............. | .................... |
| R | 2866 hCoV-19/England/MILK-B01F34/2020\|EPI_ISL_642805\|20 | .................... | .................... | ..................... | .................... | .................... | .................... |
| R | 2867 hCoV-19/England/MILK-B026A7/2020\|EPI_ISL_642806\|20 | .................... | .................... | ..................... | .................... | .................... | .................... |
| R | 2868 hCoV-19/England/MILK-B02364/2020\|EPI_ISL_642808\|20 | .................... | .................... | ..................... | .................... | .................... | .................... |
| R | 2869 hCoV-19/England/MILK-B01E82/2020\|EPI_ISL_642809\|20 | .................... | .A.................. | ..................... | .................... | .................... | ..A................. |
| R | 2870 hCoV-19/England/MILK-B028ED/2020\|EPI_ISL_642897\|20 | .................... | .................... | ..................... | .................... | .................... | .................... |
| R | 2871 hCoV-19/England/MILK-B020C7/2020\|EPI_ISL_642898\|20 | .................... | .A.................. | ..................... | .................... | .................... | ..A................. |
| R | 2872 hCoV-19/England/MILK-B02CD2/2020\|EPI_ISL_642899\|20 | .................... | .................... | ..................... | .................... | .................... | .................... |
| R | 2873 hCoV-19/Poland/PL_MCB_51/2020\|EPI_ISL_645050\|2020- | .................... | .................... | ..................... | .................... | .................... | .................... |
| R | 2874 hCoV-19/Poland/PL_MCB_52/2020\|EPI_ISL_645051\|2020- | .................... | .................... | ..................... | .................... | .................... | .................... |
| R | 2875 hCoV-19/Poland/PL_MCB_53/2020\|EPI_ISL_645052\|2020- | .................... | .................... | ..................... | .................... | .................... | .................... |
| R | 2876 hCoV-19/Poland/PL_MCB_54/2020\|EPI_ISL_645053\|2020- | .................... | .................... | ..................... | .................... | .................... | .................... |
| R | 2877 hCoV-19/Poland/PL_MCB_55/2020\|EPI_ISL_645054\|2020- | .................... | .................... | ..................... | .................... | .................... | .................... |
| R | 2878 hCoV-19/Poland/PL_MCB_56/2020\|EPI_ISL_645055\|2020- | .................... | .................... | ..................... | .................... | .................... | .................... |
| R | 2879 hCoV-19/Poland/PL_MCB_57/2020\|EPI_ISL_645056\|2020- | .................... | .................... | ..................... | .................... | .................... | .................... |
| R | 2880 hCoV-19/Poland/PL_MCB_58/2020\|EPI_ISL_645057\|2020- | .................... | .................... | ..................... | .................... | .................... | .................... |
| R | 2881 hCoV-19/Poland/PL_MCB_59/2020\|EPI_ISL_645058\|2020- | .................... | .................... | ..................... | .................... | .................... | .................... |
| R | 2882 hCoV-19/Poland/PL_MCB_60/2020\|EPI_ISL_645059\|2020- | .................... | .................... | ..................... | .................... | .................... | .................... |
| R | 2883 hCoV-19/Poland/PL_MCB_64/2020\|EPI_ISL_645063\|2020- | .................... | .................... | ..................... | .................... | .................... | .................... |
| R | 2884 hCoV-19/Poland/PL_MCB_65/2020\|EPI_ISL_645064\|2020- | .................... | .................... | ..................... | .................... | .................... | .................... |
| R | 2885 hCoV-19/Poland/PL_MCB_67/2020\|EPI_ISL_645066\|2020- | .................... | .................... | ..................... | .................... | .................... | .................... |
| R | 2886 hCoV-19/Poland/PL_MCB_71/2020\|EPI_ISL_645070\|2020- | .................... | .................... | ..................... | .................... | .................... | .................... |
| R | 2887 hCoV-19/Poland/PL_MCB_72/2020\|EPI_ISL_645071\|2020- | .................... | .................... | ..................... | .................... | .................... | .................... |
| R | 2888 hCoV-19/Poland/PL_MCB_73/2020\|EPI_ISL_645072\|2020- | .................... | .................... | ..................... | .................... | .................... | .................... |
| R | 2889 hCoV-19/Poland/PL_MCB_74/2020\|EPI_ISL_645073\|2020- | .................... | .................... | ..................... | .................... | .................... | .................... |
| R | 2890 hCoV-19/Poland/PL_MCB_75/2020\|EPI_ISL_645074\|2020- | .................... | .................... | ..................... | .................... | .................... | .................... |
| R | 2891 hCoV-19/Poland/PL_MCB_76/2020\|EPI_ISL_645075\|2020- | .................... | .................... | ..................... | .................... | .................... | .................... |
| R | 2892 hCoV-19/Poland/PL_MCB_77/2020\|EPI_ISL_645076\|2020- | .................... | .................... | ..................... | .................... | .................... | .................... |
| R | 2893 hCoV-19/Poland/PL_MCB_78/2020\|EPI_ISL_645077\|2020- | .................... | .................... | ..................... | .................... | .................... | .................... |
| R | 2894 hCoV-19/Poland/PL_MCB_79/2020\|EPI_ISL_645078\|2020- | .................... | .................... | ..................... | .................... | .................... | .................... |
| R | 2895 hCoV-19/Poland/PL_MCB_80/2020\|EPI_ISL_645079\|2020- | .................... | .................... | ..................... | .................... | .................... | .................... |
| R | 2896 hCoV-19/Poland/PL_MCB_81/2020\|EPI_ISL_645080\|2020- | .................... | .................... | ..................... | .................... | .................... | .................... |
| R | 2897 hCoV-19/Poland/PL_MCB_82/2020\|EPI_ISL_645081\|2020- | .................... | .................... | ..................... | .................... | .................... | .................... |
| R | 2898 hCoV-19/Poland/PL_MCB_83/2020\|EPI_ISL_645082\|2020- | .................... | .................... | ..................... | .................... | .................... | .................... |
| R | 2899 hCoV-19/Poland/PL_MCB_84/2020\|EPI_ISL_645083\|2020- | .................... | .................... | ..................... | .................... | .................... | .................... |
| R | 2900 hCoV-19/Poland/PL_MCB_85/2020\|EPI_ISL_645084\|2020- | .................... | .................... | ..................... | .................... | .................... | .................... |
| R | 2901 hCoV-19/Poland/PL_MCB_86/2020\|EPI_ISL_645085\|2020- | .................... | .................... | ..................... | .................... | .................... | .................... |
| R | 2902 hCoV-19/Poland/PL_MCB_87/2020\|EPI_ISL_645086\|2020- | .................... | .................... | ..................... | .................... | .................... | .................... |
| R | 2903 hCoV-19/Poland/PL_MCB_88/2020\|EPI_ISL_645087\|2020- | .................... | .................... | ..................... | .................... | .................... | .................... |
| R | 2904 hCoV-19/Poland/PL_MCB_89/2020\|EPI_ISL_645088\|2020- | .................... | .................... | ..................... | .................... | .................... | .................... |
| R | 2905 hCoV-19/Poland/PL_MCB_90/2020\|EPI_ISL_645089\|2020- | .................... | .................... | ..................... | .................... | .................... | .................... |
| R | 2906 hCoV-19/Poland/PL_MCB_101/2020\|EPI_ISL_645100\|2020 | .................... | .................... | ..................... | .................... | .................... | .................... |
| R | 2907 hCoV-19/Poland/PL_MCB_102/2020\|EPI_ISL_645101\|2020 | .................... | .................... | ..................... | .................... | .................... | .................... |
| R | 2908 hCoV-19/Poland/PL_MCB_103/2020\|EPI_ISL_645102\|2020 | .................... | .................... | ..................... | .................... | .................... | .................... |
| R | 2909 hCoV-19/Poland/PL_MCB_104/2020\|EPI_ISL_645103\|2020 | .................... | .................... | ..................... | .................... | .................... | .................... |
| R | 2910 hCoV-19/Poland/PL_MCB_105/2020\|EPI_ISL_645104\|2020 | .................... | .................... | ..................... | .................... | .................... | .................... |
| R | 2911 hCoV-19/Poland/PL_MCB_106/2020\|EPI_ISL_645105\|2020 | .................... | .................... | ..................... | .................... | .................... | .................... |
| R | 2912 hCoV-19/Poland/PL_MCB_107/2020\|EPI_ISL_645106\|2020 | .................... | .................... | ..................... | .................... | .................... | .................... |
| R | 2913 hCoV-19/Poland/PL_MCB_108/2020\|EPI_ISL_645107\|2020 | .................... | .................... | ..................... | .................... | .................... | .................... |
| R | 2914 hCoV-19/Poland/PL_MCB_109/2020\|EPI_ISL_645108\|2020 | .................... | .................... | ..................... | .................... | .................... | .................... |
| R | 2915 hCoV-19/Poland/PL_MCB_110/2020\|EPI_ISL_645109\|2020 | .................... | .................... | ..................... | .................... | .................... | .................... |
| R | 2916 hCoV-19/Singapore/1388/2020\|EPI_ISL_645115\|2020-10 | .................... | .................... | ..................... | .................... | .................... | .................... |
| R | 2917 hCoV-19/Singapore/1389/2020\|EPI_ISL_645116\|2020-09 | .................... | .................... | ..................... | .................... | .................... | .................... |
| R | 2918 hCoV-19/Singapore/1390/2020\|EPI_ISL_645117\|2020-11 | .................... | .................... | ..................... | .................... | .................... | .................... |
| R | 2919 hCoV-19/Singapore/1391/2020\|EPI_ISL_645118\|2020-11 | .................... | .................... | ..................... | .................... | .................... | .................... |
| R | 2920 hCoV-19/Singapore/1393/2020\|EPI_ISL_645120\|2020-11 | .................... | .A.................. | ..................... | .................... | .................... | ..A................. |
| R | 2921 hCoV-19/Singapore/1394/2020\|EPI_ISL_645121\|2020-11 | .................... | .A.................. | ..................... | .................... | .................... | ..A................. |
| R | 2922 hCoV-19/Singapore/1395/2020\|EPI_ISL_645122\|2020-11 | .................... | .A.................. | ..................... | .................... | .................... | ..A................. |
| R | 2923 hCoV-19/Singapore/1396/2020\|EPI_ISL_645123\|2020-11 | .................... | .A.................. | ..................... | .................... | .................... | ..A................. |
| R | 2924 hCoV-19/Singapore/1397/2020\|EPI_ISL_645124\|2020-11 | .................... | .A.................. | ..................... | .................... | .................... | ..A................. |
| R | 2925 hCoV-19/Singapore/1398/2020\|EPI_ISL_645125\|2020-11 | .................... | .A.................. | ..................... | .................... | .................... | .................... |
| R | 2926 hCoV-19/Singapore/1399/2020\|EPI_ISL_645126\|2020-11 | .................... | .A.................. | ..................... | .................... | .................... | ..A................. |
| R | 2927 hCoV-19/Singapore/1400/2020\|EPI_ISL_645127\|2020-11 | .................... | .A.................. | ..................... | .................... | .................... | ..A................. |
| R | 2928 hCoV-19/Singapore/1401/2020\|EPI_ISL_645128\|2020-11 | .................... | .A.................. | ..................... | .................... | .................... | ..A................. |
| R | 2929 hCoV-19/Singapore/1402/2020\|EPI_ISL_645129\|2020-11 | .................... | .A.................. | ..................... | .................... | .................... | .................... |
| R | 2930 hCoV-19/England/QEUH-B2E4E7/2020\|EPI_ISL_646380\|20 | .................... | .................... | ..................... | .................... | .................... | .................... |
| R | 2931 hCoV-19/England/QEUH-B31A94/2020\|EPI_ISL_646381\|20 | .................... | .................... | ..................... | .................... | .................... | .................... |
| R | 2932 hCoV-19/England/ALDP-B240CF/2020\|EPI_ISL_646382\|20 | .................... | .................... | ..................... | .................... | .................... | .................... |
| R | 2933 hCoV-19/England/QEUH-B3218F/2020\|EPI_ISL_646384\|20 | .................... | .................... | ..................... | .................... | .................... | .................... |
| R | 2934 hCoV-19/England/ALDP-B23E02/2020\|EPI_ISL_646385\|20 | .................... | .................... | ..................... | .................... | .................... | .................... |
| R | 2935 hCoV-19/England/QEUH-B2DE53/2020\|EPI_ISL_646386\|20 | .................... | .................... | ..................... | .................... | .................... | .................... |
| R | 2936 hCoV-19/England/QEUH-B2E265/2020\|EPI_ISL_646387\|20 | .................... | .................... | ..................... | .................... | .................... | .................... |
| R | 2937 hCoV-19/England/ALDP-B23FA5/2020\|EPI_ISL_646388\|20 | .................... | .................... | ..................... | .................... | .................... | .................... |
| R | 2938 hCoV-19/England/QEUH-B31A2B/2020\|EPI_ISL_646389\|20 | .................... | .................... | ..................... | .................... | .................... | .................... |

| | | | | | | | | |
|---|---|---|---|---|---|---|---|---|
| R | 2939 | hCoV-19/England/QEUH-B2E4C9/2020\|EPI_ISL_646394\|20 | ···················· | ···················· | ····················· | ···················· | ···················· | ···················· |
| R | 2940 | hCoV-19/England/QEUH-B31162/2020\|EPI_ISL_646395\|20 | ···················· | ···················· | ····················· | ···················· | ···················· | ···················· |
| R | 2941 | hCoV-19/England/QEUH-B2DE08/2020\|EPI_ISL_646396\|20 | ···················· | ···················· | ····················· | ···················· | ···················· | ···················· |
| R | 2942 | hCoV-19/England/ALDP-B23CCC/2020\|EPI_ISL_646397\|20 | ···················· | ·A·················· | ····················· | ···················· | ···················· | ··A················· |
| R | 2943 | hCoV-19/England/QEUH-B3194C/2020\|EPI_ISL_646400\|20 | ···················· | ···················· | ····················· | ···················· | ···················· | ···················· |
| R | 2944 | hCoV-19/England/QEUH-B2E3F9/2020\|EPI_ISL_646401\|20 | ···················· | ·A·················· | ····················· | ···················· | ···················· | ··A················· |
| R | 2945 | hCoV-19/England/ALDP-B23EB7/2020\|EPI_ISL_646402\|20 | ···················· | ···················· | ····················· | ···················· | ···················· | ···················· |
| R | 2946 | hCoV-19/England/QEUH-B31F77/2020\|EPI_ISL_646403\|20 | ···················· | ···················· | ····················· | ···················· | ···················· | ···················· |
| R | 2947 | hCoV-19/England/QEUH-B31CBC/2020\|EPI_ISL_646404\|20 | ···················· | ···················· | ····················· | ···················· | ···················· | ···················· |
| R | 2948 | hCoV-19/England/QEUH-B2DD65/2020\|EPI_ISL_646405\|20 | ···················· | ·A·················· | ····················· | ···················· | ···················· | ···················· |
| R | 2949 | hCoV-19/England/QEUH-B31CDA/2020\|EPI_ISL_646406\|20 | ···················· | ···················· | ····················· | ···················· | ···················· | ···················· |
| R | 2950 | hCoV-19/England/QEUH-B2E0E3/2020\|EPI_ISL_646407\|20 | ···················· | ···················· | ····················· | ···················· | ···················· | ···················· |
| R | 2951 | hCoV-19/England/QEUH-B31B91/2020\|EPI_ISL_646408\|20 | ···················· | ···················· | ····················· | ···················· | ···················· | ···················· |
| R | 2952 | hCoV-19/England/QEUH-B2F844/2020\|EPI_ISL_646409\|20 | ···················· | ···················· | ····················· | ···················· | ···················· | ···················· |
| R | 2953 | hCoV-19/England/QEUH-B2E48D/2020\|EPI_ISL_646410\|20 | ···················· | ···················· | ····················· | ···················· | ···················· | ···················· |
| R | 2954 | hCoV-19/England/QEUH-B3200A/2020\|EPI_ISL_646411\|20 | ···················· | ···················· | ····················· | ···················· | ···················· | ···················· |
| R | 2955 | hCoV-19/England/ALDP-B23E8A/2020\|EPI_ISL_646412\|20 | ···················· | ···················· | ····················· | ···················· | ···················· | ···················· |
| R | 2956 | hCoV-19/England/QEUH-B31C07/2020\|EPI_ISL_646413\|20 | ···················· | ···················· | ····················· | ···················· | ···················· | ···················· |
| R | 2957 | hCoV-19/England/QEUH-B31DE6/2020\|EPI_ISL_646414\|20 | ···················· | ···················· | ····················· | ···················· | ···················· | ···················· |
| R | 2958 | hCoV-19/England/QEUH-B31E3E/2020\|EPI_ISL_646415\|20 | ···················· | ···················· | ····················· | ···················· | ···················· | ···················· |
| R | 2959 | hCoV-19/England/QEUH-B2E195/2020\|EPI_ISL_646416\|20 | ···················· | ·A·················· | ····················· | ···················· | ···················· | ···················· |
| R | 2960 | hCoV-19/England/QEUH-B2E1B3/2020\|EPI_ISL_646417\|20 | ···················· | ·A·················· | ····················· | ···················· | ···················· | ···················· |
| R | 2961 | hCoV-19/England/ALDP-B240ED/2020\|EPI_ISL_646418\|20 | ···················· | ···················· | ····················· | ···················· | ···················· | ···················· |
| R | 2962 | hCoV-19/England/ALDP-B24083/2020\|EPI_ISL_646419\|20 | ···················· | ···················· | ····················· | ···················· | ···················· | ···················· |
| R | 2963 | hCoV-19/England/ALDP-B240A1/2020\|EPI_ISL_646420\|20 | ···················· | ···················· | ····················· | ···················· | ···················· | ···················· |
| R | 2964 | hCoV-19/England/QEUH-B310CF/2020\|EPI_ISL_646421\|20 | ···················· | ···················· | ····················· | ···················· | ······T············· | ···················· |
| R | 2965 | hCoV-19/England/QEUH-B2E05C/2020\|EPI_ISL_646422\|20 | ···················· | ···················· | ····················· | ···················· | ···················· | ···················· |
| R | 2966 | hCoV-19/England/QEUH-B2ED73/2020\|EPI_ISL_646423\|20 | ···················· | ·A·················· | ····················· | ···················· | ···················· | ···················· |
| R | 2967 | hCoV-19/England/QEUH-B2DE80/2020\|EPI_ISL_646424\|20 | ···················· | ···················· | ····················· | ···················· | ···················· | ···················· |
| R | 2968 | hCoV-19/England/QEUH-B3219E/2020\|EPI_ISL_646425\|20 | ···················· | ···················· | ····················· | ···················· | ···················· | ···················· |
| R | 2969 | hCoV-19/England/ALDP-B24214/2020\|EPI_ISL_646426\|20 | ···················· | ···················· | ····················· | ···················· | ···················· | ···················· |
| R | 2970 | hCoV-19/England/QEUH-B2DDB0/2020\|EPI_ISL_646427\|20 | ···················· | ···················· | ····················· | ···················· | ···················· | ···················· |
| R | 2971 | hCoV-19/England/ALDP-B24029/2020\|EPI_ISL_646428\|20 | ···················· | ·A·················· | ····················· | ···················· | ···················· | ···················· |
| R | 2972 | hCoV-19/England/ALDP-B23C35/2020\|EPI_ISL_646429\|20 | ···················· | ···················· | ····················· | ···················· | ···················· | ···················· |
| R | 2973 | hCoV-19/England/ALDP-B23D7E/2020\|EPI_ISL_646430\|20 | ···················· | ···················· | ····················· | ···················· | ···················· | ···················· |
| R | 2974 | hCoV-19/England/ALDP-B23E11/2020\|EPI_ISL_646431\|20 | ···················· | ······A············· | ····················· | ···················· | ···················· | ···················· |
| R | 2975 | hCoV-19/England/QEUH-B32143/2020\|EPI_ISL_646433\|20 | ···················· | ···················· | ····················· | ···················· | ···················· | ···················· |
| R | 2976 | hCoV-19/England/ALDP-B23D41/2020\|EPI_ISL_646434\|20 | ···················· | ···················· | ····················· | ···················· | ···················· | ···················· |
| R | 2977 | hCoV-19/England/QEUH-B319D3/2020\|EPI_ISL_646435\|20 | ···················· | ···················· | ····················· | ···················· | ···················· | ···················· |
| R | 2978 | hCoV-19/England/QEUH-B2E07A/2020\|EPI_ISL_646436\|20 | ···················· | ···················· | ····················· | ···················· | ···················· | ···················· |
| R | 2979 | hCoV-19/England/QEUH-B31A1C/2020\|EPI_ISL_646437\|20 | ···················· | ···················· | ····················· | ···················· | ······T············· | ···················· |
| R | 2980 | hCoV-19/England/QEUH-B31CAD/2020\|EPI_ISL_646438\|20 | ···················· | ···················· | ····················· | ···················· | ···················· | ···················· |
| R | 2981 | hCoV-19/England/QEUH-B3193D/2020\|EPI_ISL_646439\|20 | ···················· | ···················· | ····················· | ···················· | ···················· | ···················· |
| R | 2982 | hCoV-19/England/QEUH-B31A67/2020\|EPI_ISL_646440\|20 | ···················· | ···················· | ····················· | ···················· | ···················· | ···················· |
| R | 2983 | hCoV-19/England/ALDP-B24117/2020\|EPI_ISL_646441\|20 | ···················· | ···················· | ····················· | ···················· | ···················· | ···················· |
| R | 2984 | hCoV-19/England/ALDP-B23F87/2020\|EPI_ISL_646442\|20 | ···················· | ···················· | ····················· | ···················· | ···················· | ···················· |
| R | 2985 | hCoV-19/England/ALDP-B23D23/2020\|EPI_ISL_646443\|20 | ···················· | ···················· | ····················· | ···················· | ······T············· | ···················· |
| R | 2986 | hCoV-19/England/QEUH-B2F862/2020\|EPI_ISL_646444\|20 | ···················· | ···················· | ····················· | ···················· | ···················· | ···················· |
| R | 2987 | hCoV-19/England/QEUH-B318A9/2020\|EPI_ISL_646445\|20 | ···················· | ···················· | ····················· | ···················· | ···················· | ···················· |
| R | 2988 | hCoV-19/England/QEUH-B31EF2/2020\|EPI_ISL_646446\|20 | ···················· | ···················· | ····················· | ···················· | ···················· | ···················· |
| R | 2989 | hCoV-19/England/QEUH-B2ED0A/2020\|EPI_ISL_646447\|20 | ···················· | ···················· | ····················· | ···················· | ···················· | ···················· |
| R | 2990 | hCoV-19/England/ALDP-B23E6C/2020\|EPI_ISL_646448\|20 | ···················· | ···················· | ····················· | ···················· | ···················· | ···················· |
| R | 2991 | hCoV-19/England/QEUH-B31FE0/2020\|EPI_ISL_646449\|20 | ···················· | ···················· | ····················· | ···················· | ···················· | ···················· |
| R | 2992 | hCoV-19/England/QEUH-B310ED/2020\|EPI_ISL_646450\|20 | ···K················ | ·A·················· | ····················· | ···················· | ···················· | ···················· |
| R | 2993 | hCoV-19/England/QEUH-B31E6B/2020\|EPI_ISL_646452\|20 | ···················· | ···················· | ····················· | ···················· | ···················· | ···················· |
| R | 2994 | hCoV-19/England/QEUH-B2CA41/2020\|EPI_ISL_646453\|20 | ···················· | ···················· | ····················· | ···················· | ···················· | ···················· |
| R | 2995 | hCoV-19/England/ALDP-B26036/2020\|EPI_ISL_646454\|20 | ···················· | ···················· | ····················· | ···················· | ···················· | ···················· |
| R | 2996 | hCoV-19/England/QEUH-B2BAD9/2020\|EPI_ISL_646456\|20 | ···················· | ···················· | ····················· | ···················· | ···················· | ···················· |
| R | 2997 | hCoV-19/England/QEUH-B2C8A1/2020\|EPI_ISL_646457\|20 | ···················· | ···················· | ····················· | ···················· | ···················· | ···················· |
| R | 2998 | hCoV-19/England/QEUH-B2B71E/2020\|EPI_ISL_646458\|20 | ···················· | ···················· | ····················· | ···················· | ···················· | ···················· |
| R | 2999 | hCoV-19/England/QEUH-B2CA9C/2020\|EPI_ISL_646459\|20 | ···················· | ···················· | ····················· | ···················· | ···················· | ···················· |
| R | 3000 | hCoV-19/England/ALDP-B1A439/2020\|EPI_ISL_647704\|20 | ···················· | ···················· | ····················· | ···················· | ···················· | ···················· |
| R | 3001 | hCoV-19/England/ALDP-B1A7F4/2020\|EPI_ISL_647705\|20 | ···················· | ···················· | ····················· | ···················· | ···················· | ···················· |
| R | 3002 | hCoV-19/England/CAMC-B210E0/2020\|EPI_ISL_647706\|20 | ···················· | ···················· | ····················· | ···················· | ···················· | ···················· |
| R | 3003 | hCoV-19/England/CAMC-B210C2/2020\|EPI_ISL_647707\|20 | ···················· | ···················· | ····················· | ···················· | ···················· | ···················· |
| R | 3004 | hCoV-19/England/ALDP-B1AAEC/2020\|EPI_ISL_647708\|20 | ···················· | ···················· | ····················· | ···················· | ···················· | ···················· |
| R | 3005 | hCoV-19/England/CAMC-B20597/2020\|EPI_ISL_647709\|20 | ···················· | ·A·················· | ····················· | ···················· | ···················· | ···················· |
| R | 3006 | hCoV-19/England/CAMC-B1FDB1/2020\|EPI_ISL_647710\|20 | ···················· | ···················· | ····················· | ···················· | ······T············· | ···············A···· |
| R | 3007 | hCoV-19/England/ALDP-B1AAA0/2020\|EPI_ISL_647711\|20 | ···················· | ···················· | ····················· | ···················· | ···················· | ···················· |
| R | 3008 | hCoV-19/England/CAMC-B20A5C/2020\|EPI_ISL_647713\|20 | ···················· | ···················· | ····················· | ···················· | ···················· | ···················· |
| R | 3009 | hCoV-19/England/CAMC-B20F6C/2020\|EPI_ISL_647714\|20 | ···················· | ···················· | ····················· | ···················· | ···················· | ···················· |
| R | 3010 | hCoV-19/England/CAMC-B20BB3/2020\|EPI_ISL_647715\|20 | ···················· | ···················· | ····················· | ···················· | ···················· | ···················· |
| R | 3011 | hCoV-19/England/CAMC-B1FF06/2020\|EPI_ISL_647716\|20 | ···················· | ···················· | ····················· | ···················· | ···················· | ···················· |
| R | 3012 | hCoV-19/England/CAMC-B20A2F/2020\|EPI_ISL_647717\|20 | ···················· | ···················· | ····················· | ···················· | ···················· | ···················· |
| R | 3013 | hCoV-19/England/CAMC-B20B0E/2020\|EPI_ISL_647718\|20 | ···················· | ···················· | ····················· | ···················· | ···················· | ···················· |
| R | 3014 | hCoV-19/England/CAMC-B20DCC/2020\|EPI_ISL_647719\|20 | ···················· | ···················· | ····················· | ···················· | ···················· | ···················· |
| R | 3015 | hCoV-19/England/CAMC-B20F11/2020\|EPI_ISL_647721\|20 | ···················· | ···················· | ····················· | ···················· | ···················· | ···················· |
| R | 3016 | hCoV-19/England/CAMC-B1FEFA/2020\|EPI_ISL_647722\|20 | ···················· | ···················· | ····················· | ···················· | ···················· | ···················· |
| R | 3017 | hCoV-19/England/CAMC-B20C47/2020\|EPI_ISL_647724\|20 | ···················· | ···················· | ····················· | ···················· | ···················· | ···················· |
| R | 3018 | hCoV-19/England/CAMC-B20AE3/2020\|EPI_ISL_647725\|20 | ···················· | ···················· | ····················· | ···················· | ···················· | ···················· |

| | | | | | | | |
|---|---|---|---|---|---|---|---|
| R | 3019 hCoV-19/England/CAMC-B1FEDC/2020\|EPI_ISL_647727\|20 | ···················· | ···················· | ···················· | ···················· | ···················· | ···················· |
| R | 3020 hCoV-19/England/CAMC-B1FD84/2020\|EPI_ISL_647728\|20 | ···················· | ···················· | ···················· | ···················· | ···················· | ···················· |
| R | 3021 hCoV-19/England/CAMC-B20C0B/2020\|EPI_ISL_647729\|20 | ···················· | ···················· | ···················· | ···················· | ···················· | ···················· |
| R | 3022 hCoV-19/England/CAMC-B20A3E/2020\|EPI_ISL_647731\|20 | ···················· | ···················· | ···················· | ···················· | ···················· | ···················· |
| R | 3023 hCoV-19/England/CAMC-B20F7B/2020\|EPI_ISL_647732\|20 | ···················· | ···················· | ···················· | ···················· | ···················· | ···················· |
| R | 3024 hCoV-19/England/ALDP-B1A81F/2020\|EPI_ISL_647733\|20 | ···················· | ···················· | ···················· | ···················· | ···················· | ···················· |
| R | 3025 hCoV-19/England/CAMC-B212EA/2020\|EPI_ISL_647734\|20 | ···················· | ···················· | ···················· | ···················· | ···················· | ···················· |
| R | 3026 hCoV-19/England/CAMC-B20AF2/2020\|EPI_ISL_647735\|20 | ···················· | ·A·················· | ···················· | ···················· | ···················· | ···A················ |
| R | 3027 hCoV-19/England/CAMC-B20E8D/2020\|EPI_ISL_647736\|20 | ···················· | ···················· | ···················· | ···················· | ···················· | ···················· |
| R | 3028 hCoV-19/England/CAMC-B205C4/2020\|EPI_ISL_647737\|20 | ···················· | ···················· | ···················· | ···················· | ···················· | ···················· |
| R | 3029 hCoV-19/England/CAMC-B1FE18/2020\|EPI_ISL_647738\|20 | ···················· | ···················· | ···················· | ···················· | ···················· | ···················· |
| R | 3030 hCoV-19/England/CAMC-B20AA7/2020\|EPI_ISL_647739\|20 | ···················· | ···················· | ···················· | ···················· | ···················· | ···················· |
| R | 3031 hCoV-19/England/CAMC-B204A9/2020\|EPI_ISL_647740\|20 | ···················· | ···················· | ···················· | ···················· | ···················· | ···················· |
| R | 3032 hCoV-19/England/ALDP-B1A457/2020\|EPI_ISL_647741\|20 | ···················· | ···················· | ···················· | ···················· | ···················· | ···················· |
| R | 3033 hCoV-19/England/CAMC-B1FB6C/2020\|EPI_ISL_647742\|20 | ···················· | ···················· | ···················· | ···················· | ······T············· | ···················· |
| R | 3034 hCoV-19/England/CAMC-B1FCF0/2020\|EPI_ISL_647743\|20 | ···················· | ···················· | ···················· | ···················· | ···················· | ···················· |
| R | 3035 hCoV-19/England/CAMC-B20EC9/2020\|EPI_ISL_647744\|20 | ···················· | ···················· | ···················· | ···················· | ···················· | ···················· |
| R | 3036 hCoV-19/England/CAMC-B2097D/2020\|EPI_ISL_647747\|20 | ···················· | ···················· | ···················· | ···················· | ···················· | ···················· |
| R | 3037 hCoV-19/England/CAMC-B20FA8/2020\|EPI_ISL_647748\|20 | ···················· | ·A·················· | ···················· | ···················· | ···················· | ···A················ |
| R | 3038 hCoV-19/England/ALDP-B1A9D0/2020\|EPI_ISL_647750\|20 | ···················· | ···················· | ···················· | ···················· | ···················· | ···················· |
| R | 3039 hCoV-19/England/CAMC-B20D71/2020\|EPI_ISL_647751\|20 | ···················· | ···················· | ···················· | ···················· | ······T············· | ···················· |
| R | 3040 hCoV-19/England/ALDP-B1AB61/2020\|EPI_ISL_647752\|20 | ···················· | ···················· | ···················· | ···················· | ···················· | ···················· |
| R | 3041 hCoV-19/England/CAMC-B20931/2020\|EPI_ISL_647753\|20 | ···················· | ···················· | ···················· | ···················· | ···················· | ···················· |
| R | 3042 hCoV-19/England/CAMC-B20BC2/2020\|EPI_ISL_647755\|20 | ···················· | ···················· | ···················· | ···················· | ···················· | ···················· |
| R | 3043 hCoV-19/England/CAMC-B1FE27/2020\|EPI_ISL_647756\|20 | ···················· | ·A·················· | ···················· | ···················· | ···················· | ···A················ |
| R | 3044 hCoV-19/England/CAMC-B1FB99/2020\|EPI_ISL_647757\|20 | ···················· | ···················· | ···················· | ···················· | ···················· | ···················· |
| R | 3045 hCoV-19/England/CAMC-B20F4E/2020\|EPI_ISL_647758\|20 | ···················· | ···················· | ···················· | ···················· | ···················· | ···················· |
| R | 3046 hCoV-19/England/CAMC-B2046D/2020\|EPI_ISL_647759\|20 | ···················· | ···················· | ···················· | ···················· | ···················· | ···················· |
| R | 3047 hCoV-19/England/CAMC-B20719/2020\|EPI_ISL_647760\|20 | ···················· | ···················· | ···················· | ···················· | ···················· | ···················· |
| R | 3048 hCoV-19/England/CAMC-B20807/2020\|EPI_ISL_647761\|20 | ···················· | ···················· | ···················· | ···················· | ···················· | ···················· |
| R | 3049 hCoV-19/England/ALDP-B1A527/2020\|EPI_ISL_647762\|20 | ···················· | ···················· | ···················· | ···················· | ···················· | ···················· |
| R | 3050 hCoV-19/England/CAMC-B20A4D/2020\|EPI_ISL_647763\|20 | ···················· | ···················· | ···················· | ···················· | ···················· | ···················· |
| R | 3051 hCoV-19/England/CAMC-B1FDA2/2020\|EPI_ISL_647764\|20 | ···················· | ···················· | ···················· | ···················· | ···················· | ···················· |
| R | 3052 hCoV-19/England/CAMC-B2099B/2020\|EPI_ISL_647765\|20 | ···················· | ···················· | ···················· | ···················· | ···················· | ···················· |
| R | 3053 hCoV-19/England/CAMC-B20B4A/2020\|EPI_ISL_647766\|20 | ···················· | ···················· | ···················· | ···················· | ···················· | ···················· |
| R | 3054 hCoV-19/England/CAMC-B212AE/2020\|EPI_ISL_647767\|20 | ···················· | ···················· | ···················· | ···················· | ···················· | ···················· |
| R | 3055 hCoV-19/England/CAMC-B20272/2020\|EPI_ISL_647769\|20 | ···················· | ···················· | ···················· | ···················· | ···················· | ···················· |
| R | 3056 hCoV-19/England/CAMC-B1FBC6/2020\|EPI_ISL_647770\|20 | ···················· | ···················· | ···················· | ···················· | ······T············· | ···················· |
| R | 3057 hCoV-19/England/ALDP-B1A8F1/2020\|EPI_ISL_647771\|20 | ···················· | ···················· | ···················· | ···················· | ···················· | ···················· |
| R | 3058 hCoV-19/England/CAMC-B206EF/2020\|EPI_ISL_647772\|20 | ···················· | ···················· | ···················· | ···················· | ···················· | ···················· |
| R | 3059 hCoV-19/England/CAMC-B20C74/2020\|EPI_ISL_647774\|20 | ···················· | ···················· | ···················· | ···················· | ···················· | ···················· |
| R | 3060 hCoV-19/England/CAMC-B210D1/2020\|EPI_ISL_647775\|20 | ···················· | ···················· | ···················· | ···················· | ···················· | ···················· |
| R | 3061 hCoV-19/England/ALDP-B1A9FE/2020\|EPI_ISL_647776\|20 | ···················· | ·A·················· | ···················· | ···················· | ···················· | ···················· |
| R | 3062 hCoV-19/England/CAMC-B1FBB7/2020\|EPI_ISL_647777\|20 | ···················· | ···················· | ···················· | ···················· | ···················· | ···················· |
| R | 3063 hCoV-19/England/CAMC-B21244/2020\|EPI_ISL_647778\|20 | ···················· | ···················· | ···················· | ···················· | ···················· | ···················· |
| R | 3064 hCoV-19/England/CAMC-B1FE09/2020\|EPI_ISL_647779\|20 | ···················· | ···················· | ···················· | ···················· | ···················· | ···················· |
| R | 3065 hCoV-19/England/CAMC-B211DE/2020\|EPI_ISL_647782\|20 | ···················· | ···················· | ···················· | ···················· | ···················· | ···················· |
| R | 3066 hCoV-19/England/CAMC-B1FCE1/2020\|EPI_ISL_647783\|20 | ···················· | ···················· | ···················· | ···················· | ···················· | ···················· |
| R | 3067 hCoV-19/England/CAMC-B209D7/2020\|EPI_ISL_647784\|20 | ···················· | ···················· | ···················· | ···················· | ···················· | ···················· |
| R | 3068 hCoV-19/England/CAMC-B20C65/2020\|EPI_ISL_647785\|20 | ···················· | ···················· | ···················· | ···················· | ···················· | ···················· |
| R | 3069 hCoV-19/England/ALDP-B1A4FD/2020\|EPI_ISL_647786\|20 | ···················· | ···················· | ···················· | ···················· | ···················· | ···················· |
| R | 3070 hCoV-19/England/ALDP-B1A4A2/2020\|EPI_ISL_647787\|20 | ···················· | ···················· | ···················· | ···················· | ···················· | ···················· |
| R | 3071 hCoV-19/England/CAMC-B20B3B/2020\|EPI_ISL_647788\|20 | ···················· | ·A·················· | ···················· | ···················· | ···················· | ···················· |
| R | 3072 hCoV-19/England/CAMC-B1FF60/2020\|EPI_ISL_647789\|20 | ···················· | ···················· | ···················· | ···················· | ···················· | ···················· |
| R | 3079 hCoV-19/USA/MI-UM-10036578381/2020\|EPI_ISL_640212\| | ···················· | ······A············· | ···················· | ···················· | ···················· | ···················· |
| R | 3081 hCoV-19/USA/MI-UM-10036599140/2020\|EPI_ISL_640215\| | ···················· | ·A·················· | ···················· | ···················· | ···················· | ···················· |
| R | 3084 hCoV-19/Germany/BW-MVZ-LS-01/2020\|EPI_ISL_640219\|2 | ···················· | ···················· | ···················· | ···················· | ···················· | ···················· |
| R | 3085 hCoV-19/Germany/BW-MVZ-LS-02/2020\|EPI_ISL_640220\|2 | ···················· | ···················· | ···················· | ···················· | ···················· | ···················· |
| R | 3086 hCoV-19/Germany/BW-MVZ-LS-03/2020\|EPI_ISL_640221\|2 | ···················· | ···················· | ···················· | ···················· | ···················· | ···················· |
| R | 3087 hCoV-19/Germany/BW-MVZ-LS-04/2020\|EPI_ISL_640222\|2 | ···················· | ···················· | ···················· | ···················· | ···················· | ···················· |
| R | 3088 hCoV-19/Germany/BW-MVZ-LS-05/2020\|EPI_ISL_640223\|2 | ···················· | ···················· | ···················· | ···················· | ···················· | ···················· |
| R | 3089 hCoV-19/Germany/BW-MVZ-LS-06/2020\|EPI_ISL_640224\|2 | ···················· | ···················· | ···················· | ···················· | ···················· | ···················· |
| R | 3090 hCoV-19/Germany/BW-MVZ-LS-07/2020\|EPI_ISL_640225\|2 | ···················· | ···················· | ···················· | ···················· | ···················· | ···················· |
| R | 3091 hCoV-19/Germany/BW-MVZ-LS-08/2020\|EPI_ISL_640226\|2 | ···················· | ···················· | ···················· | ···················· | ···················· | ···················· |
| R | 3092 hCoV-19/Germany/BW-MVZ-LS-09/2020\|EPI_ISL_640227\|2 | ···················· | ···················· | ···················· | ···················· | ···················· | ···················· |
| R | 3093 hCoV-19/Germany/BW-MVZ-LS-10/2020\|EPI_ISL_640228\|2 | ···················· | ···················· | ···················· | ···················· | ···················· | ···················· |
| R | 3094 hCoV-19/Germany/BW-MVZ-LS-11/2020\|EPI_ISL_640229\|2 | ···················· | ···················· | ···················· | ···················· | ···················· | ···················· |
| R | 3095 hCoV-19/Germany/BW-MVZ-LS-13/2020\|EPI_ISL_640231\|2 | ···················· | ···················· | ···················· | ···················· | ···················· | ···················· |
| R | 3096 hCoV-19/Germany/BW-MVZ-LS-14/2020\|EPI_ISL_640232\|2 | ···················· | ······A············· | ···················· | ···················· | ···················· | ···················· |
| R | 3097 hCoV-19/Germany/BW-MVZ-LS-15/2020\|EPI_ISL_640233\|2 | ···················· | ···················· | ···················· | ···················· | ···················· | ···················· |
| R | 3098 hCoV-19/mink/Denmark/mDK-152/2020\|EPI_ISL_641516\|2 | ···················· | ·N·················· | ···················· | ···················· | ···················· | ···················· |
| R | 3099 hCoV-19/France/PAC-SC235/2020\|EPI_ISL_641520\|2020- | ···················· | ···················· | ···················· | ···················· | ···················· | ···················· |
| R | 3100 hCoV-19/France/PAC-SC236/2020\|EPI_ISL_641522\|2020- | ···················· | ···················· | ···················· | ···················· | ···················· | ···················· |
| R | 3101 hCoV-19/France/PAC-SC237/2020\|EPI_ISL_641523\|2020- | ···················· | ···················· | ···················· | ···················· | ···················· | ···················· |
| R | 3102 hCoV-19/France/PAC-SC238/2020\|EPI_ISL_641524\|2020- | ···················· | ···················· | ···················· | ···················· | ···················· | ···················· |
| R | 3103 hCoV-19/France/PAC-SC239/2020\|EPI_ISL_641525\|2020- | ···················· | ···················· | ···················· | ···················· | ···················· | ···················· |
| R | 3104 hCoV-19/France/PAC-SC240/2020\|EPI_ISL_641526\|2020- | ···················· | ···················· | ···················· | ···················· | ···················· | ···················· |
| R | 3105 hCoV-19/France/PAC-SC242/2020\|EPI_ISL_641527\|2020- | ···················· | ···················· | ···················· | ···················· | ···················· | ···················· |
| R | 3106 hCoV-19/France/PAC-SC243/2020\|EPI_ISL_641528\|2020- | ···················· | ···················· | ···················· | ···················· | ···················· | ···················· |
| R | 3107 hCoV-19/France/PAC-SC244/2020\|EPI_ISL_641529\|2020- | ···················· | ···················· | ···················· | ···················· | ···················· | ···················· |

| | | | | | | | |
|---|---|---|---|---|---|---|---|
| R | 3108 hCoV-19/France/PAC-SC245/2020\|EPI_ISL_641530\|2020- | ···················· | ···················· | ····················· | ···················· | ···················· | ···················· |
| R | 3109 hCoV-19/France/PAC-SC246/2020\|EPI_ISL_641531\|2020- | ···················· | ···················· | ····················· | ···················· | ···················· | ···················· |
| R | 3110 hCoV-19/France/PAC-SC247/2020\|EPI_ISL_641532\|2020- | ···················· | ···················· | ····················· | ···················· | ···················· | ···················· |
| R | 3111 hCoV-19/France/PAC-SC248/2020\|EPI_ISL_641533\|2020- | ···················· | ···················· | ····················· | ···················· | ···················· | ···················· |
| R | 3112 hCoV-19/France/PAC-SC249/2020\|EPI_ISL_641534\|2020- | ···················· | ···················· | ····················· | ···················· | ···················· | ···················· |
| R | 3113 hCoV-19/France/ARA-SC310/2020\|EPI_ISL_641535\|2020- | ···················· | ···················· | ····················· | ···················· | ···················· | ···················· |
| R | 3114 hCoV-19/France/ARA-SC311/2020\|EPI_ISL_641536\|2020- | ···················· | ···················· | ····················· | ···················· | ···················· | ···················· |
| R | 3115 hCoV-19/France/ARA-SC314/2020\|EPI_ISL_641537\|2020- | ···················· | ···················· | ····················· | ···················· | ···················· | ···················· |
| R | 3116 hCoV-19/France/ARA-SC319/2020\|EPI_ISL_641538\|2020- | ···················· | ···················· | ····················· | ···················· | ···················· | ···················· |
| R | 3117 hCoV-19/France/ARA-SC322/2020\|EPI_ISL_641539\|2020- | ···················· | ···················· | ····················· | ···················· | ···················· | ···················· |
| R | 3118 hCoV-19/France/ARA-SC324/2020\|EPI_ISL_641540\|2020- | ···················· | ···················· | ····················· | ···················· | ···················· | ···················· |
| R | 3119 hCoV-19/France/ARA-SC333/2020\|EPI_ISL_641541\|2020- | ···················· | ···················· | ····················· | ···················· | ···················· | ···················· |
| R | 3120 hCoV-19/France/ARA-SC340/2020\|EPI_ISL_641542\|2020- | ···················· | ···················· | ····················· | ···················· | ···················· | ···················· |
| R | 3121 hCoV-19/France/ARA-SC346/2020\|EPI_ISL_641543\|2020- | ···················· | ···················· | ····················· | ···················· | ···················· | ···················· |
| R | 3122 hCoV-19/France/ARA-SC350/2020\|EPI_ISL_641544\|2020- | ···················· | ···················· | ····················· | ···················· | ···················· | ···················· |
| R | 3123 hCoV-19/France/ARA-SC359/2020\|EPI_ISL_641545\|2020- | ···················· | ···················· | ····················· | ···················· | ···················· | ···················· |
| R | 3124 hCoV-19/France/ARA-SC397/2020\|EPI_ISL_641546\|2020- | ···················· | ···················· | ····················· | ···················· | ···················· | ···················· |
| R | 3125 hCoV-19/France/ARA-SC401/2020\|EPI_ISL_641547\|2020- | ···················· | ···················· | ····················· | ···················· | ···················· | ···················· |
| R | 3126 hCoV-19/France/ARA-SC404/2020\|EPI_ISL_641548\|2020- | ···················· | ···················· | ····················· | ···················· | ···················· | ···················· |
| R | 3127 hCoV-19/France/ARA-SC405/2020\|EPI_ISL_641549\|2020- | ···················· | ···················· | ····················· | ···················· | ···················· | ···················· |
| R | 3128 hCoV-19/France/ARA-SC413/2020\|EPI_ISL_641550\|2020- | ···················· | ···················· | ···········T········· | ···················· | ···················· | ···················· |
| R | 3129 hCoV-19/France/OCC-SC415/2020\|EPI_ISL_641551\|2020- | ···················· | ···················· | ····················· | ···················· | ···················· | ···················· |
| R | 3130 hCoV-19/France/OCC-SC417/2020\|EPI_ISL_641552\|2020- | ···················· | ···················· | ····················· | ···················· | ···················· | ···················· |
| R | 3131 hCoV-19/France/OCC-SC418/2020\|EPI_ISL_641553\|2020- | ···················· | ···················· | ····················· | ···················· | ···················· | ···················· |
| R | 3132 hCoV-19/France/OCC-SC427/2020\|EPI_ISL_641554\|2020- | ···················· | ···················· | ····················· | ···················· | ···················· | ···················· |
| R | 3133 hCoV-19/France/OCC-SC432/2020\|EPI_ISL_641555\|2020- | ···················· | ···················· | ····················· | ···················· | ···················· | ···················· |
| R | 3134 hCoV-19/France/OCC-SC465/2020\|EPI_ISL_641556\|2020- | ···················· | ···················· | ····················· | ···················· | ···················· | ···················· |
| R | 3135 hCoV-19/Belgium/ULG-10715/2020\|EPI_ISL_641557\|2020 | ···················· | ···················· | ····················· | ···················· | ···················· | ···················· |
| R | 3142 hCoV-19/Belgium/ULG-10722/2020\|EPI_ISL_641564\|2020 | ···················· | ···················· | ····················· | ···················· | ···················· | ···················· |
| R | 3144 hCoV-19/England/MILK-B060C3/2020\|EPI_ISL_642810\|20 | ···················· | ···················· | ····················· | ···················· | ···················· | ···················· |
| R | 3145 hCoV-19/England/MILK-B02A14/2020\|EPI_ISL_642811\|20 | ···················· | ···················· | ····················· | ···················· | ···················· | ···················· |
| R | 3146 hCoV-19/England/MILK-B024F8/2020\|EPI_ISL_642812\|20 | ···················· | ···················· | ····················· | ···················· | ···················· | ···················· |
| R | 3147 hCoV-19/England/MILK-B02C96/2020\|EPI_ISL_642814\|20 | ···················· | ···················· | ····················· | ···················· | ···················· | ···················· |
| R | 3148 hCoV-19/England/MILK-B05D81/2020\|EPI_ISL_642815\|20 | ···················· | ···················· | ····················· | ···················· | ···················· | ···················· |
| R | 3149 hCoV-19/England/MILK-B0603C/2020\|EPI_ISL_642816\|20 | ···················· | ···················· | ····················· | ···················· | ···················· | ···················· |
| R | 3150 hCoV-19/England/MILK-B02012/2020\|EPI_ISL_642817\|20 | ···················· | ···················· | ····················· | ···················· | ···················· | ···················· |
| R | 3151 hCoV-19/England/MILK-B060E1/2020\|EPI_ISL_642818\|20 | ···················· | ···················· | ····················· | ···················· | ···················· | ···················· |
| R | 3152 hCoV-19/England/MILK-B05DFA/2020\|EPI_ISL_642831\|20 | ···················· | ·A·················· | ····················· | ···················· | ···················· | ···················· |
| R | 3153 hCoV-19/England/MILK-B05F9A/2020\|EPI_ISL_642832\|20 | ···················· | ···················· | ····················· | ···················· | ······T············· | ···················· |
| R | 3154 hCoV-19/England/MILK-B0605A/2020\|EPI_ISL_642834\|20 | ···················· | ···················· | ····················· | ···················· | ···················· | ···················· |
| R | 3155 hCoV-19/England/MILK-B0273B/2020\|EPI_ISL_642835\|20 | ···················· | ···················· | ····················· | ···················· | ···················· | ···················· |
| R | 3156 hCoV-19/England/MILK-B01F16/2020\|EPI_ISL_642837\|20 | ···················· | ···················· | ····················· | ···················· | ···················· | ···················· |
| R | 3157 hCoV-19/England/MILK-B026E3/2020\|EPI_ISL_642838\|20 | ···················· | ···················· | ····················· | ···················· | ···················· | ···················· |
| R | 3158 hCoV-19/England/MILK-B06263/2020\|EPI_ISL_642839\|20 | ···················· | ···················· | ····················· | ···················· | ···················· | ···················· |
| R | 3159 hCoV-19/England/MILK-B02E09/2020\|EPI_ISL_642840\|20 | ···················· | ···················· | ····················· | ···················· | ···················· | ···················· |
| R | 3160 hCoV-19/England/MILK-B05E42/2020\|EPI_ISL_642841\|20 | ···················· | ···················· | ····················· | ···················· | ···················· | ···················· |
| R | 3161 hCoV-19/England/MILK-B01F25/2020\|EPI_ISL_642842\|20 | ···················· | ···················· | ····················· | ···················· | ···················· | ···················· |
| R | 3162 hCoV-19/England/MILK-B01DB2/2020\|EPI_ISL_642843\|20 | ···················· | ···················· | ····················· | ···················· | ···················· | ···················· |
| R | 3163 hCoV-19/England/MILK-B021B5/2020\|EPI_ISL_642844\|20 | ···················· | ···················· | ····················· | ···················· | ···················· | ···················· |
| R | 3164 hCoV-19/England/MILK-B0638E/2020\|EPI_ISL_642845\|20 | ···················· | ·A·················· | ····················· | ···················· | ···················· | ···················· |
| R | 3165 hCoV-19/England/MILK-B02874/2020\|EPI_ISL_642846\|20 | ···················· | ···················· | ····················· | ···················· | ···················· | ···················· |
| R | 3166 hCoV-19/England/MILK-B02BB7/2020\|EPI_ISL_642847\|20 | ···················· | ···················· | ····················· | ···················· | ···················· | ···················· |
| R | 3167 hCoV-19/England/MILK-B02BF3/2020\|EPI_ISL_642848\|20 | ···················· | ···················· | ····················· | ···················· | ···················· | ···················· |
| R | 3168 hCoV-19/England/MILK-B02D1B/2020\|EPI_ISL_642849\|20 | ···················· | ···················· | ····················· | ···················· | ···················· | ···················· |
| R | 3169 hCoV-19/England/MILK-B02883/2020\|EPI_ISL_642850\|20 | ···················· | ···················· | ····················· | ···················· | ···················· | ···················· |
| R | 3170 hCoV-19/England/MILK-B031A5/2020\|EPI_ISL_642851\|20 | ···················· | ···················· | ····················· | ···················· | ···················· | ···················· |
| R | 3171 hCoV-19/England/MILK-B062EB/2020\|EPI_ISL_642854\|20 | ···················· | ···················· | ····················· | ···················· | ···················· | ···················· |
| R | 3172 hCoV-19/England/MILK-B06157/2020\|EPI_ISL_642855\|20 | ···················· | ···················· | ····················· | ···················· | ···················· | ···················· |
| R | 3173 hCoV-19/England/MILK-B0274A/2020\|EPI_ISL_642856\|20 | ···················· | ···················· | ····················· | ···················· | ···················· | ···················· |
| R | 3174 hCoV-19/England/MILK-B0266B/2020\|EPI_ISL_642857\|20 | ···················· | ·A·················· | ····················· | ···················· | ···················· | ···A················ |
| R | 3175 hCoV-19/England/MILK-B0263E/2020\|EPI_ISL_642858\|20 | ···················· | ···················· | ····················· | ···················· | ···················· | ···················· |
| R | 3176 hCoV-19/England/MILK-B02795/2020\|EPI_ISL_642859\|20 | ···················· | ···················· | ····················· | ···················· | ···················· | ···················· |
| R | 3177 hCoV-19/England/MILK-B026B6/2020\|EPI_ISL_642860\|20 | ···················· | ···················· | ····················· | ···················· | ···················· | ···················· |
| R | 3178 hCoV-19/England/MILK-B05F12/2020\|EPI_ISL_642861\|20 | ···················· | ···················· | ····················· | ···················· | ···················· | ···················· |
| R | 3179 hCoV-19/England/MILK-B060A5/2020\|EPI_ISL_642862\|20 | ···················· | ···················· | ····················· | ···················· | ···················· | ···················· |
| R | 3180 hCoV-19/England/MILK-B028FC/2020\|EPI_ISL_642863\|20 | ···················· | ···················· | ····················· | ···················· | ···················· | ···················· |
| R | 3181 hCoV-19/England/MILK-B0264D/2020\|EPI_ISL_642864\|20 | ···················· | ···················· | ····················· | ···················· | ···················· | ···················· |
| R | 3182 hCoV-19/England/MILK-B02926/2020\|EPI_ISL_642865\|20 | ···················· | ···················· | ····················· | ···················· | ······T············· | ···················· |
| R | 3183 hCoV-19/England/MILK-B05E06/2020\|EPI_ISL_642866\|20 | ···················· | ···················· | ····················· | ···················· | ···················· | ···················· |
| R | 3184 hCoV-19/England/MILK-B02188/2020\|EPI_ISL_642867\|20 | ···················· | ···················· | ····················· | ···················· | ···················· | ···················· |
| R | 3185 hCoV-19/England/MILK-B02CE1/2020\|EPI_ISL_642869\|20 | ···················· | ···················· | ····················· | ···················· | ···················· | ···················· |
| R | 3186 hCoV-19/Wales/MILK-B02C2D/2020\|EPI_ISL_642871\|2020 | ···················· | ···················· | ····················· | ···················· | ···················· | ···················· |
| R | 3187 hCoV-19/England/MILK-B027FF/2020\|EPI_ISL_642872\|20 | ···················· | ···················· | ····················· | ···················· | ···················· | ···················· |
| R | 3188 hCoV-19/England/MILK-B01E19/2020\|EPI_ISL_642873\|20 | ···················· | ···················· | ····················· | ···················· | ···················· | ···················· |
| R | 3189 hCoV-19/England/MILK-B02100/2020\|EPI_ISL_642874\|20 | ···················· | ···················· | ····················· | ···················· | ···················· | ···················· |
| R | 3190 hCoV-19/England/MILK-B0299F/2020\|EPI_ISL_642876\|20 | ···················· | ···················· | ····················· | ···················· | ······T············· | ···················· |
| R | 3191 hCoV-19/England/MILK-B02434/2020\|EPI_ISL_642877\|20 | ···················· | ···················· | ····················· | ···················· | ···················· | ···················· |
| R | 3192 hCoV-19/England/MILK-B02425/2020\|EPI_ISL_642878\|20 | ···················· | ···················· | ····················· | ···················· | ···················· | ···················· |
| R | 3193 hCoV-19/England/MILK-B028DE/2020\|EPI_ISL_642879\|20 | ···················· | ···················· | ····················· | ···················· | ···················· | ···················· |
| R | 3194 hCoV-19/England/MILK-B023FB/2020\|EPI_ISL_642880\|20 | ···················· | ···················· | ····················· | ···················· | ···················· | ···················· |

| | | | | | | | |
|---|---|---|---|---|---|---|---|
| R | 3195 hCoV-19/England/MILK-B02B20/2020\|EPI_ISL_642883\|20 | ···················· | ···················· | ···················· | ···················· | ···················· | ···················· |
| R | 3196 hCoV-19/England/MILK-B0270E/2020\|EPI_ISL_642884\|20 | ···················· | ···················· | ···················· | ···················· | ···················· | ···················· |
| R | 3197 hCoV-19/England/MILK-B05DAF/2020\|EPI_ISL_642885\|20 | ···················· | ·A·················· | ···················· | ···················· | ···················· | ··A················· |
| R | 3198 hCoV-19/England/MILK-B01D49/2020\|EPI_ISL_642886\|20 | ···················· | ···················· | ···················· | ···················· | ···················· | ···················· |
| R | 3199 hCoV-19/England/MILK-B02AD8/2020\|EPI_ISL_642887\|20 | ···················· | ···················· | ···················· | ···················· | ···················· | ···················· |
| R | 3200 hCoV-19/England/MILK-B02D66/2020\|EPI_ISL_642888\|20 | ···················· | ···················· | ···················· | ···················· | ···················· | ···················· |
| R | 3201 hCoV-19/England/MILK-B05EAC/2020\|EPI_ISL_642889\|20 | ···················· | ···················· | ···················· | ···················· | ···················· | ···················· |
| R | 3202 hCoV-19/England/MILK-B01E91/2020\|EPI_ISL_642890\|20 | ···················· | ···················· | ···················· | ···················· | ···················· | ···················· |
| R | 3203 hCoV-19/England/MILK-B02971/2020\|EPI_ISL_642891\|20 | ···················· | ···················· | ···················· | ···················· | ·····T·············· | ···················· |
| R | 3204 hCoV-19/England/MILK-B02D75/2020\|EPI_ISL_642892\|20 | ···················· | ···················· | ···················· | ···················· | ···················· | ···················· |
| R | 3205 hCoV-19/England/MILK-B01EBF/2020\|EPI_ISL_642893\|20 | ···················· | ···················· | ···················· | ···················· | ···················· | ···················· |
| R | 3206 hCoV-19/England/MILK-B02610/2020\|EPI_ISL_642894\|20 | ···················· | ···················· | ···················· | ···················· | ·····T·············· | ···················· |
| R | 3207 hCoV-19/England/MILK-B0602D/2020\|EPI_ISL_642895\|20 | ···················· | ···················· | ···················· | ···················· | ·····T·············· | ···················· |
| R | 3209 hCoV-19/Poland/PL_MCB_10/2020\|EPI_ISL_645009\|2020- | ···················· | ···················· | ···················· | ···················· | ···················· | ···················· |
| R | 3210 hCoV-19/Poland/PL_MCB_11/2020\|EPI_ISL_645010\|2020- | ···················· | ···················· | ···················· | ···················· | ···················· | ···················· |
| R | 3211 hCoV-19/Poland/PL_MCB_12/2020\|EPI_ISL_645011\|2020- | ···················· | ···················· | ···················· | ···················· | ···················· | ···················· |
| R | 3212 hCoV-19/Poland/PL_MCB_13/2020\|EPI_ISL_645012\|2020- | ···················· | ···················· | ···················· | ···················· | ···················· | ···················· |
| R | 3213 hCoV-19/Poland/PL_MCB_14/2020\|EPI_ISL_645013\|2020- | ···················· | ···················· | ···················· | ···················· | ···················· | ···················· |
| R | 3214 hCoV-19/Poland/PL_MCB_15/2020\|EPI_ISL_645014\|2020- | ···················· | ···················· | ···················· | ···················· | ···················· | ···················· |
| R | 3215 hCoV-19/Poland/PL_MCB_16/2020\|EPI_ISL_645015\|2020- | ···················· | ···················· | ···················· | ···················· | ···················· | ···················· |
| R | 3216 hCoV-19/Poland/PL_MCB_17/2020\|EPI_ISL_645016\|2020- | ···················· | ···················· | ···················· | ···················· | ···················· | ···················· |
| R | 3217 hCoV-19/Poland/PL_MCB_18/2020\|EPI_ISL_645017\|2020- | ···················· | ···················· | ···················· | ···················· | ···················· | ···················· |
| R | 3218 hCoV-19/Poland/PL_MCB_19/2020\|EPI_ISL_645018\|2020- | ···················· | ···················· | ···················· | ···················· | ···················· | ···················· |
| R | 3219 hCoV-19/Poland/PL_MCB_20/2020\|EPI_ISL_645019\|2020- | ···················· | ···················· | ···················· | ···················· | ···················· | ···················· |
| R | 3220 hCoV-19/Poland/PL_MCB_21/2020\|EPI_ISL_645020\|2020- | ···················· | ···················· | ···················· | ···················· | ···················· | ···················· |
| R | 3221 hCoV-19/Poland/PL_MCB_22/2020\|EPI_ISL_645021\|2020- | ···················· | ···················· | ···················· | ···················· | ···················· | ···················· |
| R | 3222 hCoV-19/Poland/PL_MCB_23/2020\|EPI_ISL_645022\|2020- | ···················· | ···················· | ···················· | ···················· | ···················· | ···················· |
| R | 3223 hCoV-19/Poland/PL_MCB_24/2020\|EPI_ISL_645023\|2020- | ···················· | ···················· | ···················· | ···················· | ···················· | ···················· |
| R | 3224 hCoV-19/Poland/PL_MCB_25/2020\|EPI_ISL_645024\|2020- | ···················· | ···················· | ···················· | ···················· | ···················· | ···················· |
| R | 3225 hCoV-19/Poland/PL_MCB_26/2020\|EPI_ISL_645025\|2020- | ···················· | ·A·················· | ···················· | ···················· | ···················· | ···················· |
| R | 3226 hCoV-19/Poland/PL_MCB_27/2020\|EPI_ISL_645026\|2020- | ···················· | ·A·················· | ···················· | ···················· | ···················· | ···················· |
| R | 3227 hCoV-19/Poland/PL_MCB_28/2020\|EPI_ISL_645027\|2020- | ···················· | ···················· | ···················· | ···················· | ···················· | ···················· |
| R | 3228 hCoV-19/Poland/PL_MCB_29/2020\|EPI_ISL_645028\|2020- | ···················· | ···················· | ···················· | ···················· | ···················· | ···················· |
| R | 3229 hCoV-19/Poland/PL_MCB_30/2020\|EPI_ISL_645029\|2020- | ···················· | ···················· | ···················· | ···················· | ···················· | ···················· |
| R | 3230 hCoV-19/Poland/PL_MCB_31/2020\|EPI_ISL_645030\|2020- | ···················· | ···················· | ···················· | ···················· | ···················· | ···················· |
| R | 3231 hCoV-19/Poland/PL_MCB_32/2020\|EPI_ISL_645031\|2020- | ···················· | ···················· | ···················· | ···················· | ···················· | ···················· |
| R | 3232 hCoV-19/Poland/PL_MCB_33/2020\|EPI_ISL_645032\|2020- | ···················· | ···················· | ···················· | ···················· | ···················· | ···················· |
| R | 3233 hCoV-19/Poland/PL_MCB_34/2020\|EPI_ISL_645033\|2020- | ···················· | ···················· | ···················· | ···················· | ···················· | ···················· |
| R | 3234 hCoV-19/Poland/PL_MCB_35/2020\|EPI_ISL_645034\|2020- | ···················· | ···················· | ···················· | ···················· | ···················· | ···················· |
| R | 3235 hCoV-19/Poland/PL_MCB_36/2020\|EPI_ISL_645035\|2020- | ···················· | ···················· | ···················· | ···················· | ···················· | ···················· |
| R | 3236 hCoV-19/Poland/PL_MCB_37/2020\|EPI_ISL_645036\|2020- | ···················· | ···················· | ···················· | ···················· | ···················· | ···················· |
| R | 3237 hCoV-19/Poland/PL_MCB_38/2020\|EPI_ISL_645037\|2020- | ···················· | ···················· | ···················· | ···················· | ···················· | ···················· |
| R | 3238 hCoV-19/Poland/PL_MCB_39/2020\|EPI_ISL_645038\|2020- | ···················· | ···················· | ···················· | ···················· | ···················· | ···················· |
| R | 3239 hCoV-19/Poland/PL_MCB_40/2020\|EPI_ISL_645039\|2020- | ···················· | ···················· | ···················· | ···················· | ···················· | ···················· |
| R | 3240 hCoV-19/Poland/PL_MCB_41/2020\|EPI_ISL_645040\|2020- | ···················· | ···················· | ···················· | ···················· | ···················· | ···················· |
| R | 3241 hCoV-19/Poland/PL_MCB_42/2020\|EPI_ISL_645041\|2020- | ···················· | ···················· | ···················· | ···················· | ···················· | ···················· |
| R | 3242 hCoV-19/Poland/PL_MCB_43/2020\|EPI_ISL_645042\|2020- | ···················· | ···················· | ···················· | ···················· | ···················· | ···················· |
| R | 3243 hCoV-19/Poland/PL_MCB_44/2020\|EPI_ISL_645043\|2020- | ···················· | ·A·················· | ···················· | ···················· | ···················· | ···················· |
| R | 3244 hCoV-19/Poland/PL_MCB_45/2020\|EPI_ISL_645044\|2020- | ···················· | ···················· | ···················· | ···················· | ···················· | ···················· |
| R | 3245 hCoV-19/Poland/PL_MCB_46/2020\|EPI_ISL_645045\|2020- | ···················· | ···················· | ···················· | ···················· | ···················· | ···················· |
| R | 3246 hCoV-19/Poland/PL_MCB_47/2020\|EPI_ISL_645046\|2020- | ···················· | ···················· | ···················· | ···················· | ···················· | ···················· |
| R | 3247 hCoV-19/Poland/PL_MCB_48/2020\|EPI_ISL_645047\|2020- | ···················· | ···················· | ···················· | ···················· | ···················· | ···················· |
| R | 3248 hCoV-19/Poland/PL_MCB_49/2020\|EPI_ISL_645048\|2020- | ···················· | ···················· | ···················· | ···················· | ···················· | ···················· |
| R | 3249 hCoV-19/Poland/PL_MCB_50/2020\|EPI_ISL_645049\|2020- | ···················· | ···················· | ···················· | ···················· | ···················· | ···················· |
| R | 3250 hCoV-19/Poland/PL_MCB_61/2020\|EPI_ISL_645060\|2020- | ···················· | ···················· | ···················· | ···················· | ···················· | ···················· |
| R | 3251 hCoV-19/Poland/PL_MCB_62/2020\|EPI_ISL_645061\|2020- | ···················· | ···················· | ···················· | ···················· | ···················· | ···················· |
| R | 3252 hCoV-19/Poland/PL_MCB_63/2020\|EPI_ISL_645062\|2020- | ···················· | ···················· | ···················· | ···················· | ···················· | ···················· |
| R | 3253 hCoV-19/Poland/PL_MCB_66/2020\|EPI_ISL_645065\|2020- | ···················· | ···················· | ···················· | ···················· | ···················· | ···················· |
| R | 3254 hCoV-19/Poland/PL_MCB_68/2020\|EPI_ISL_645067\|2020- | ···················· | ···················· | ···················· | ···················· | ···················· | ···················· |
| R | 3255 hCoV-19/Poland/PL_MCB_69/2020\|EPI_ISL_645068\|2020- | ···················· | ···················· | ···················· | ···················· | ···················· | ···················· |
| R | 3256 hCoV-19/Poland/PL_MCB_70/2020\|EPI_ISL_645069\|2020- | ···················· | ···················· | ···················· | ···················· | ···················· | ···················· |
| R | 3257 hCoV-19/Poland/PL_MCB_91/2020\|EPI_ISL_645090\|2020- | ···················· | ···················· | ···················· | ···················· | ···················· | ···················· |
| R | 3258 hCoV-19/Poland/PL_MCB_92/2020\|EPI_ISL_645091\|2020- | ···················· | ···················· | ···················· | ···················· | ···················· | ···················· |
| R | 3259 hCoV-19/England/QEUH-B31F59/2020\|EPI_ISL_646340\|20 | ···················· | ···················· | ···················· | ···················· | ···················· | ···················· |
| R | 3260 hCoV-19/England/QEUH-B31DC8/2020\|EPI_ISL_646341\|20 | ···················· | ···················· | ···················· | ···················· | ···················· | ···················· |
| R | 3261 hCoV-19/England/ALDP-B23BED/2020\|EPI_ISL_646342\|20 | ···················· | ···················· | ···················· | ···················· | ···················· | ···················· |
| R | 3262 hCoV-19/England/ALDP-B23E20/2020\|EPI_ISL_646343\|20 | ···················· | ···················· | ···················· | ···················· | ···················· | ···················· |
| R | 3263 hCoV-19/England/QEUH-B2DDED/2020\|EPI_ISL_646344\|20 | ···················· | ···················· | ···················· | ···················· | ···················· | ···················· |
| R | 3264 hCoV-19/England/ALDP-B24065/2020\|EPI_ISL_646345\|20 | ···················· | ···················· | ···················· | ···················· | ···················· | ···················· |
| R | 3265 hCoV-19/England/ALDP-B23E3F/2020\|EPI_ISL_646346\|20 | ···················· | ···················· | ···················· | ···················· | ···················· | ···················· |
| R | 3266 hCoV-19/England/ALDP-B23FD2/2020\|EPI_ISL_646347\|20 | ···················· | ···················· | ···················· | ···················· | ···················· | ···················· |
| R | 3267 hCoV-19/England/QEUH-B320FB/2020\|EPI_ISL_646348\|20 | ···················· | ···················· | ···················· | ···················· | ···················· | ···················· |
| R | 3268 hCoV-19/England/ALDP-B23F69/2020\|EPI_ISL_646349\|20 | ···················· | ···················· | ···················· | ···················· | ···················· | ···················· |
| R | 3269 hCoV-19/England/ALDP-B23EC6/2020\|EPI_ISL_646350\|20 | ···················· | ···················· | ···················· | ···················· | ···················· | ···················· |
| R | 3270 hCoV-19/England/QEUH-B2E47E/2020\|EPI_ISL_646351\|20 | ···················· | ···················· | ···················· | ···················· | ···················· | ···················· |
| R | 3271 hCoV-19/England/QEUH-B31997/2020\|EPI_ISL_646352\|20 | ···················· | ···················· | ···················· | ···················· | ···················· | ···················· |
| R | 3272 hCoV-19/England/QEUH-B319F1/2020\|EPI_ISL_646353\|20 | ···················· | ·A·················· | ···················· | ···················· | ···················· | ···················· |
| R | 3273 hCoV-19/England/ALDP-B23FB4/2020\|EPI_ISL_646354\|20 | ···················· | ···················· | ···················· | ···················· | ···················· | ···················· |
| R | 3274 hCoV-19/England/QEUH-B2ECD0/2020\|EPI_ISL_646355\|20 | ···················· | ·A·················· | ···················· | ···················· | ···················· | ···················· |
| R | 3275 hCoV-19/England/ALDP-B24108/2020\|EPI_ISL_646356\|20 | ···················· | ···················· | ···················· | ···················· | ···················· | ···················· |

```
R  3276 hCoV-19/England/QEUH-B2DD83/2020|EPI_ISL_646357|20   ····················  ····················  ····················  ····················  ····················  ····················
R  3277 hCoV-19/England/QEUH-B2E20B/2020|EPI_ISL_646358|20   ····················  ····················  ····················  ····················  ······T·············  ····················
R  3278 hCoV-19/England/QEUH-B2DDFC/2020|EPI_ISL_646359|20   ····················  ····················  ····················  ····················  ····················  ····················
R  3279 hCoV-19/England/QEUH-B31C34/2020|EPI_ISL_646360|20   ····················  ····················  ····················  ····················  ····················  ····················
R  3280 hCoV-19/England/QEUH-B31AEF/2020|EPI_ISL_646361|20   ····················  ····················  ····················  ····················  ····················  ····················
R  3281 hCoV-19/England/ALDP-B241BD/2020|EPI_ISL_646362|20   ····················  ····················  ····················  ····················  ····················  ····················
R  3282 hCoV-19/England/ALDP-B23F1E/2020|EPI_ISL_646363|20   ····················  ····················  ····················  ····················  ····················  ····················
R  3283 hCoV-19/England/QEUH-B310A1/2020|EPI_ISL_646364|20   ····················  ····················  ····················  ····················  ····················  ····················
R  3284 hCoV-19/England/QEUH-B2DE9F/2020|EPI_ISL_646365|20   ····················  ····················  ····················  ····················  ····················  ····················
R  3285 hCoV-19/England/ALDP-B23FF0/2020|EPI_ISL_646366|20   ····················  ····················  ····················  ····················  ····················  ····················
R  3286 hCoV-19/England/QEUH-B2E177/2020|EPI_ISL_646368|20   ····················  ····················  ····················  ····················  ····················  ····················
R  3287 hCoV-19/England/ALDP-B23B56/2020|EPI_ISL_646369|20   ····················  ····················  ····················  ····················  ····················  ····················
R  3288 hCoV-19/England/ALDP-B23DE7/2020|EPI_ISL_646370|20   ····················  ····················  ····················  ····················  ····················  ····················
R  3289 hCoV-19/England/QEUH-B2ECEF/2020|EPI_ISL_646371|20   ····················  ····················  ····················  ····················  ····················  ····················
R  3290 hCoV-19/England/QEUH-B2E4BA/2020|EPI_ISL_646372|20   ····················  ····················  ····················  ····················  ····················  ····················
R  3291 hCoV-19/England/ALDP-B23C53/2020|EPI_ISL_646373|20   ····················  ····················  ····················  ····················  ····················  ····················
R  3292 hCoV-19/England/QEUH-B3119F/2020|EPI_ISL_646374|20   ····················  ····················  ····················  ····················  ····················  ····················
R  3293 hCoV-19/England/ALDP-B23F0F/2020|EPI_ISL_646375|20   ····················  ····················  ····················  ····················  ····················  ····················
R  3294 hCoV-19/England/ALDP-B23D6F/2020|EPI_ISL_646376|20   ····················  ····················  ····················  ····················  ····················  ····················
R  3295 hCoV-19/England/ALDP-B23D32/2020|EPI_ISL_646377|20   ····················  ····················  ····················  ····················  ····················  ····················
R  3296 hCoV-19/England/QEUH-B31D5F/2020|EPI_ISL_646378|20   ····················  ····················  ····················  ····················  ····················  ····················
R  3297 hCoV-19/England/QEUH-B31D9B/2020|EPI_ISL_646379|20   ····················  ····················  ····················  ····················  ····················  ····················
R  3298 hCoV-19/England/QEUH-B31144/2020|EPI_ISL_646390|20   ····················  ····················  ····················  ····················  ····················  ····················
R  3299 hCoV-19/England/QEUH-B2DE44/2020|EPI_ISL_646391|20   ····················  ····················  ····················  ····················  ····················  ····················
R  3300 hCoV-19/England/QEUH-B32037/2020|EPI_ISL_646392|20   ····················  ····················  ····················  ····················  ····················  ····················
R  3301 hCoV-19/England/QEUH-B31F95/2020|EPI_ISL_646399|20   ····················  ····················  ····················  ····················  ····················  ····················
R  3302 hCoV-19/England/CAMC-B1FCB4/2020|EPI_ISL_647670|20   ····················  ····················  ····················  ····················  ····················  ····················
R  3303 hCoV-19/England/CAMC-B20C83/2020|EPI_ISL_647671|20   ····················  ····················  ····················  ····················  ····················  ····················
R  3304 hCoV-19/England/ALDP-B1A8B5/2020|EPI_ISL_647672|20   ····················  ····················  ····················  ····················  ····················  ····················
R  3305 hCoV-19/England/CAMC-B20E23/2020|EPI_ISL_647673|20   ····················  ····················  ····················  ····················  ····················  ····················
R  3306 hCoV-19/England/CAMC-B1FEEB/2020|EPI_ISL_647674|20   ····················  ····················  ····················  ····················  ····················  ····················
R  3307 hCoV-19/England/CAMC-B20D08/2020|EPI_ISL_647675|20   ····················  ····················  ····················  ····················  ····················  ····················
R  3308 hCoV-19/England/CAMC-B20EAB/2020|EPI_ISL_647676|20   ····················  ····················  ····················  ····················  ····················  ····················
R  3309 hCoV-19/England/ALDP-B1A9A3/2020|EPI_ISL_647677|20   ····················  ····················  ····················  ····················  ····················  ····················
R  3310 hCoV-19/England/CAMC-B20A89/2020|EPI_ISL_647679|20   ····················  ····················  ····················  ····················  ····················  ····················
R  3311 hCoV-19/England/CAMC-B20B1D/2020|EPI_ISL_647681|20   ····················  ····················  ····················  ····················  ······T·············  ····················
R  3312 hCoV-19/England/ALDP-B1A590/2020|EPI_ISL_647682|20   ····················  ····················  ····················  ····················  ······T·············  ····················
R  3313 hCoV-19/England/ALDP-B1AADD/2020|EPI_ISL_647683|20   ····················  ····················  ····················  ····················  ····················  ····················
R  3314 hCoV-19/England/CAMC-B1FBE4/2020|EPI_ISL_647684|20   ····················  ····················  ····················  ····················  ····················  ····················
R  3315 hCoV-19/England/CAMC-B212DB/2020|EPI_ISL_647685|20   ····················  ····················  ····················  ····················  ····················  ····················
R  3316 hCoV-19/England/CAMC-B1FBD5/2020|EPI_ISL_647686|20   ····················  ·A··················  ····················  ····················  ····················  ··A·················
R  3317 hCoV-19/England/CAMC-B20C56/2020|EPI_ISL_647687|20   ····················  ····················  ····················  ····················  ····················  ····················
R  3318 hCoV-19/England/CAMC-B21262/2020|EPI_ISL_647688|20   ····················  ····················  ····················  ····················  ····················  ····················
R  3319 hCoV-19/England/CAMC-B1FD1B/2020|EPI_ISL_647690|20   ····················  ····················  ····················  ····················  ····················  ····················
R  3320 hCoV-19/England/CAMC-B20D62/2020|EPI_ISL_647691|20   ····················  ····················  ····················  ····················  ······T·············  ····················
R  3321 hCoV-19/England/CAMC-B20DEA/2020|EPI_ISL_647692|20   ····················  ····················  ····················  ····················  ····················  ····················
R  3322 hCoV-19/England/ALDP-B1AA82/2020|EPI_ISL_647693|20   ····················  ····················  ····················  ····················  ····················  ····················
R  3323 hCoV-19/England/CAMC-B20852/2020|EPI_ISL_647694|20   ····················  ····················  ····················  ····················  ····················  ····················
R  3324 hCoV-19/England/ALDP-B1A475/2020|EPI_ISL_647695|20   ····················  ····················  ····················  ····················  ····················  ····················
R  3325 hCoV-19/England/CAMC-B20C29/2020|EPI_ISL_647696|20   ····················  ····················  ····················  ····················  ····················  ····················
R  3326 hCoV-19/England/CAMC-B2096E/2020|EPI_ISL_647697|20   ····················  ····················  ····················  ····················  ······T·············  ····················
R  3327 hCoV-19/England/ALDP-B1A888/2020|EPI_ISL_647698|20   ····················  ····················  ····················  ····················  ····················  ····················
R  3328 hCoV-19/England/CAMC-B1FD0C/2020|EPI_ISL_647699|20   ····················  ····················  ····················  ····················  ····················  ····················
R  3329 hCoV-19/South Africa/NHLS-UCT-GS-1855/2020|EPI_ISL   ····················  ·A··················  ····················  ····················  ····················  ····················
R  3330 hCoV-19/South Africa/NHLS-UCT-GS-1856/2020|EPI_ISL   ····················  ·A··················  ····················  ····················  ····················  ····················
R  3331 hCoV-19/South Africa/NHLS-UCT-GS-1874/2020|EPI_ISL   ····················  ·A··················  ····················  ····················  ····················  ····················
R  3332 hCoV-19/South Africa/NHLS-UCT-GS-2028/2020|EPI_ISL   ····················  ·A··················  ····················  ····················  ····················  ····················
R  3333 hCoV-19/South Africa/NHLS-UCT-GS-2030/2020|EPI_ISL   ····················  ·A··················  ····················  ····················  ····················  ····················
R  3334 hCoV-19/South Africa/NHLS-UCT-GS-2031/2020|EPI_ISL   ····················  ·A··················  ····················  ····················  ····················  ····················
R  3335 hCoV-19/South Africa/NHLS-UCT-GS-2032/2020|EPI_ISL   ····················  ····················  ····················  ····················  ····················  ····················
R  3336 hCoV-19/South Africa/NHLS-UCT-GS-2033/2020|EPI_ISL   ····················  ·A··················  ····················  ····················  ····················  ····················
R  3337 hCoV-19/South Africa/NHLS-UCT-GS-2083/2020|EPI_ISL   ····················  ····················  ····················  ····················  ····················  ····················
R  3338 hCoV-19/South Africa/NHLS-UCT-GS-2085/2020|EPI_ISL   ····················  ····················  ····················  ····················  ····················  ····················
R  3339 hCoV-19/South Africa/NHLS-UCT-GS-2087/2020|EPI_ISL   ····················  ····················  ····················  ····················  ····················  ····················
R  3340 hCoV-19/South Africa/NHLS-UCT-GS-2089/2020|EPI_ISL   ····················  ····················  ····················  ····················  ····················  ····················
R  3341 hCoV-19/South Africa/NHLS-UCT-GS-3461/2020|EPI_ISL   ····················  ·A··················  ····················  ····················  ····················  ····················
R  3342 hCoV-19/South Africa/NHLS-UCT-GS-3462/2020|EPI_ISL   ····················  ····················  ····················  ····················  ····················  ····················
R  3343 hCoV-19/South Africa/NHLS-UCT-GS-3468/2020|EPI_ISL   ····················  ····················  ····················  ····················  ····················  ····················
R  3344 hCoV-19/South Africa/NHLS-UCT-GS-3469/2020|EPI_ISL   ····················  ····················  ····················  ····················  ····················  ····················
R  3345 hCoV-19/South Africa/NHLS-UCT-GS-3470/2020|EPI_ISL   ····················  ····················  ····················  ····················  ····················  ····················
R  3346 hCoV-19/South Africa/NHLS-UCT-GS-3472/2020|EPI_ISL   ····················  ·A··················  ····················  ····················  ····················  ····················
R  3347 hCoV-19/South Africa/NHLS-UCT-GS-3516/2020|EPI_ISL   ····················  ·A··················  ····················  ····················  ····················  ····················
R  3348 hCoV-19/South Africa/NHLS-UCT-GS-3522/2020|EPI_ISL   ····················  ····················  ····················  ····················  ····················  ····················
R  3349 hCoV-19/USA/MI-UM-10036535783/2020|EPI_ISL_640143|   ····················  ·············A······  ····················  ····················  ····················  ····················
R  3350 hCoV-19/USA/MI-UM-10036535366/2020|EPI_ISL_640144|   ····················  ····················  ····················  ····················  ····················  ····················
R  3351 hCoV-19/USA/MI-UM-10036540707/2020|EPI_ISL_640145|   ····················  ·A··················  ····················  ····················  ····················  ····················
R  3355 hCoV-19/USA/MI-UM-10036539730/2020|EPI_ISL_640149|   ····················  ····················  ····················  ····················  ····················  ····················
R  3356 hCoV-19/USA/MI-UM-10036535538/2020|EPI_ISL_640150|   ····················  ····················  ····················  ····················  ····················  ····················
R  3357 hCoV-19/USA/MI-UM-10036530731/2020|EPI_ISL_640151|   ····················  ····················  ····················  ····················  ····················  ····················
R  3358 hCoV-19/USA/MI-UM-10036535367/2020|EPI_ISL_640152|   ····················  ····················  ····················  ····················  ····················  ····················
```

R 3360 hCoV-19/USA/MI-UM-10036540841/2020|EPI_ISL_640154| ···················· ···················· ····················· ···················· ···················· ····················
R 3361 hCoV-19/USA/MI-UM-10036531598/2020|EPI_ISL_640155| ···················· ···················· ····················· ···················· ···················· ····················
R 3362 hCoV-19/USA/MI-UM-10036537564/2020|EPI_ISL_640156| ···················· ···················· ····················· ···················· ···················· ····················
R 3363 hCoV-19/USA/MI-UM-10036535827/2020|EPI_ISL_640157| ···················· ···················· ····················· ···················· ···················· ····················
R 3364 hCoV-19/USA/MI-UM-10036579570/2020|EPI_ISL_640158| ···················· ···················· ····················· ···················· ···················· ····················
R 3365 hCoV-19/USA/MI-UM-10036541568/2020|EPI_ISL_640160| ···················· ···················· ····················· ···················· ···················· ····················
R 3366 hCoV-19/USA/MI-UM-10036424364/2020|EPI_ISL_640161| ···················· ·A·················· ····················· ···················· ···················· ····················
R 3367 hCoV-19/USA/MI-UM-10036532868/2020|EPI_ISL_640162| ···················· ···················· ····················· ···················· ···················· ····················
R 3370 hCoV-19/USA/MI-UM-10036554773/2020|EPI_ISL_640165| ···················· ···················· ····················· ···················· ···················· ····················
R 3371 hCoV-19/USA/MI-UM-10036557522/2020|EPI_ISL_640166| ···················· ···················· ····················· ···················· ···················· ····················
R 3372 hCoV-19/USA/MI-UM-10036576559/2020|EPI_ISL_640167| ···················· ······A············· ····················· ···················· ···················· ····················
R 3373 hCoV-19/USA/MI-UM-10036578598/2020|EPI_ISL_640169| ···················· ···················· ····················· ···················· ···················· ····················
R 3374 hCoV-19/USA/MI-UM-10036561032/2020|EPI_ISL_640170| ···················· ···················· ····················· ···················· ···················· ····················
R 3375 hCoV-19/USA/MI-UM-10036552359/2020|EPI_ISL_640171| ···················· ···················· ····················· ···················· ···················· ····················
R 3376 hCoV-19/USA/MI-UM-10036554113/2020|EPI_ISL_640172| ···················· ···················· ····················· ···················· ···················· ····················
R 3377 hCoV-19/USA/MI-UM-10036580137/2020|EPI_ISL_640174| ···················· ······A············· ····················· ···················· ···················· ····················
R 3378 hCoV-19/USA/MI-UM-10036548314/2020|EPI_ISL_640175| ···················· ···················· ····················· ···················· ···················· ····················
R 3379 hCoV-19/USA/MI-UM-10036561341/2020|EPI_ISL_640176| ···················· ···················· ····················· ···················· ···················· ····················
R 3380 hCoV-19/USA/MI-UM-10036571011/2020|EPI_ISL_640177| ···················· ···················· ····················· ···················· ···················· ····················
R 3381 hCoV-19/USA/MI-UM-10036554756/2020|EPI_ISL_640178| ···················· ···················· ····················· ···················· ···················· ····················
R 3382 hCoV-19/USA/MI-UM-10036561998/2020|EPI_ISL_640179| ···················· ···················· ····················· ···················· ···················· ····················
R 3383 hCoV-19/USA/MI-UM-10036561091/2020|EPI_ISL_640180| ···················· ···················· ····················· ···················· ···················· ····················
R 3384 hCoV-19/USA/MI-UM-10036555954/2020|EPI_ISL_640181| ···················· ···················· ····················· ···················· ···················· ····················
R 3390 hCoV-19/USA/MI-UM-10036577639/2020|EPI_ISL_640190| ···················· ···················· ····················· ···················· ···················· ····················
R 3391 hCoV-19/USA/MI-UM-10036556049/2020|EPI_ISL_640191| ···················· ···················· ····················· ···················· ···················· ····················
R 3396 hCoV-19/USA/MI-UM-10036575544/2020|EPI_ISL_640198| ···················· ···················· ····················· ···················· ···················· ····················
R 3397 hCoV-19/England/MILK-B022B2/2020|EPI_ISL_642776|20 ···················· ···················· ····················· ···················· ···················· ····················
R 3398 hCoV-19/England/MILK-B05FB8/2020|EPI_ISL_642777|20 ···················· ···················· ····················· ···················· ···················· ····················
R 3399 hCoV-19/England/MILK-B02319/2020|EPI_ISL_642778|20 ···················· ···················· ····················· ···················· ···················· ····················
R 3400 hCoV-19/England/MILK-B01EDD/2020|EPI_ISL_642780|20 ···················· ···················· ····················· ···················· ···················· ····················
R 3401 hCoV-19/England/MILK-B01CD3/2020|EPI_ISL_642781|20 ···················· ···················· ····················· ···················· ···················· ····················
R 3402 hCoV-19/England/MILK-B01EFB/2020|EPI_ISL_642782|20 ···················· ···················· ····················· ···················· ···················· ····················
R 3403 hCoV-19/England/MILK-B01DD0/2020|EPI_ISL_642783|20 ···················· ···················· ····················· ···················· ···················· ····················
R 3404 hCoV-19/England/MILK-B06087/2020|EPI_ISL_642784|20 ···················· ···················· ····················· ···················· ···················· ····················
R 3405 hCoV-19/England/MILK-B062CD/2020|EPI_ISL_642785|20 ···················· ···················· ····················· ···················· ···················· ····················
R 3406 hCoV-19/England/MILK-B061A2/2020|EPI_ISL_642786|20 ···················· ···················· ····················· ···················· ······T············· ···············A····
R 3407 hCoV-19/England/MILK-B029EA/2020|EPI_ISL_642787|20 ···················· ···················· ····················· ···················· ···················· ····················
R 3408 hCoV-19/England/MILK-B02258/2020|EPI_ISL_642788|20 ···················· ···················· ····················· ···················· ···················· ····················
R 3409 hCoV-19/England/MILK-B060F0/2020|EPI_ISL_642789|20 ···················· ···················· ····················· ···················· ···················· ····················
R 3410 hCoV-19/England/MILK-B02328/2020|EPI_ISL_642790|20 ···················· ···················· ····················· ···················· ···················· ····················
R 3411 hCoV-19/England/MILK-B0281A/2020|EPI_ISL_642791|20 ···················· ···················· ····················· ···················· ···················· ····················
R 3412 hCoV-19/England/MILK-B02AC9/2020|EPI_ISL_642792|20 ···················· ···················· ····················· ···················· ······T············· ····················
R 3413 hCoV-19/England/MILK-B05F6D/2020|EPI_ISL_642793|20 ···················· ···················· ····················· ···················· ···················· ····················
R 3414 hCoV-19/England/MILK-B0208B/2020|EPI_ISL_642795|20 ···················· ···················· ····················· ···················· ···················· ····················
R 3415 hCoV-19/England/MILK-B029CC/2020|EPI_ISL_642796|20 ···················· ···················· ····················· ···················· ···················· ····················
R 3416 hCoV-19/England/MILK-B05E7F/2020|EPI_ISL_642797|20 ···················· ···················· ····················· ···················· ···················· ····················
R 3417 hCoV-19/England/MILK-B02838/2020|EPI_ISL_642798|20 ···················· ···················· ····················· ···················· ···················· ····················
R 3418 hCoV-19/England/MILK-B01E0A/2020|EPI_ISL_642799|20 ···················· ···················· ····················· ···················· ···················· ····················
R 3419 hCoV-19/England/MILK-B0205E/2020|EPI_ISL_642819|20 ···················· ···················· ····················· ···················· ······T············· ····················
R 3420 hCoV-19/England/MILK-B02A05/2020|EPI_ISL_642821|20 ···················· ···················· ····················· ···················· ···················· ····················
R 3421 hCoV-19/England/MILK-B0216A/2020|EPI_ISL_642822|20 ···················· ···················· ····················· ···················· ···················· ····················
R 3422 hCoV-19/England/MILK-B0600F/2020|EPI_ISL_642823|20 ···················· ···················· ····················· ···················· ···················· ····················
R 3423 hCoV-19/England/MILK-B0257D/2020|EPI_ISL_642824|20 ···················· ···················· ····················· ···················· ···················· ····················
R 3424 hCoV-19/England/MILK-B020F4/2020|EPI_ISL_642825|20 ···················· ···················· ····················· ···················· ···················· ····················
R 3425 hCoV-19/England/MILK-B02AAB/2020|EPI_ISL_642827|20 ···················· ···················· ····················· ···················· ···················· ····················
R 3426 hCoV-19/England/MILK-B06315/2020|EPI_ISL_642828|20 ···················· ···················· ····················· ···················· ···················· ····················
R 3427 hCoV-19/England/MILK-B023CE/2020|EPI_ISL_642829|20 ···················· ···················· ····················· ···················· ······T············· ····················
R 3428 hCoV-19/England/MILK-B02D2A/2020|EPI_ISL_642830|20 ···················· ···················· ····················· ···················· ···················· ····················
R 3429 hCoV-19/Poland/PL_MCB_1/2020|EPI_ISL_645000|2020-0 ···················· ···················· ····················· ···················· ···················· ····················
R 3430 hCoV-19/Poland/PL_MCB_2/2020|EPI_ISL_645001|2020-0 ···················· ···················· ····················· ···················· ···················· ····················
R 3431 hCoV-19/Poland/PL_MCB_3/2020|EPI_ISL_645002|2020-0 ···················· ···················· ····················· ···················· ···················· ····················
R 3432 hCoV-19/Poland/PL_MCB_4/2020|EPI_ISL_645003|2020-0 ···················· ···················· ····················· ···················· ···················· ····················
R 3433 hCoV-19/Poland/PL_MCB_5/2020|EPI_ISL_645004|2020-0 ···················· ···················· ····················· ···················· ···················· ····················
R 3434 hCoV-19/Poland/PL_MCB_6/2020|EPI_ISL_645005|2020-0 ···················· ···················· ····················· ···················· ···················· ····················
R 3435 hCoV-19/Poland/PL_MCB_7/2020|EPI_ISL_645006|2020-0 ···················· ···················· ····················· ···················· ···················· ····················
R 3436 hCoV-19/Poland/PL_MCB_8/2020|EPI_ISL_645007|2020-0 ···················· ···················· ····················· ···················· ···················· ····················
R 3437 hCoV-19/Poland/PL_MCB_9/2020|EPI_ISL_645008|2020-0 ···················· ···················· ····················· ···················· ···················· ····················
R 3438 hCoV-19/Poland/PL_MCB_93/2020|EPI_ISL_645092|2020- ···················· ···················· ····················· ···················· ···················· ····················
R 3439 hCoV-19/Poland/PL_MCB_94/2020|EPI_ISL_645093|2020- ···················· ···················· ····················· ···················· ···················· ····················
R 3440 hCoV-19/Poland/PL_MCB_95/2020|EPI_ISL_645094|2020- ···················· ···················· ····················· ···················· ···················· ····················
R 3441 hCoV-19/Poland/PL_MCB_96/2020|EPI_ISL_645095|2020- ···················· ···················· ····················· ···················· ···················· ····················
R 3442 hCoV-19/Poland/PL_MCB_97/2020|EPI_ISL_645096|2020- ···················· ···················· ····················· ···················· ···················· ····················
R 3443 hCoV-19/Poland/PL_MCB_98/2020|EPI_ISL_645097|2020- ···················· ···················· ····················· ···················· ···················· ····················
R 3444 hCoV-19/Poland/PL_MCB_99/2020|EPI_ISL_645098|2020- ···················· ···················· ····················· ···················· ···················· ····················
R 3445 hCoV-19/Poland/PL_MCB_100/2020|EPI_ISL_645099|2020 ···················· ···················· ····················· ···················· ···················· ····················
R 3446 hCoV-19/England/QEUH-B2DD92/2020|EPI_ISL_646300|20 ···················· ···················· ····················· ···················· ···················· ····················
R 3447 hCoV-19/England/QEUH-B31F1D/2020|EPI_ISL_646301|20 ···················· ···················· ····················· ···················· ···················· ····················
R 3448 hCoV-19/England/QEUH-B318D6/2020|EPI_ISL_646302|20 ···················· ···················· ····················· ···················· ···················· ····················
R 3449 hCoV-19/England/ALDP-B23DBA/2020|EPI_ISL_646303|20 ···················· ···················· ····················· ···················· ···················· ····················
R 3450 hCoV-19/England/QEUH-B32055/2020|EPI_ISL_646304|20 ···················· ···················· ····················· ···················· ···················· ····················

| | | | | | | | |
|---|---|---|---|---|---|---|---|
| R | 3451 hCoV-19/England/ALDP-B23EF3/2020\|EPI_ISL_646305\|20 | ···················· | ···················· | ····················· | ···················· | ···················· | ···················· |
| R | 3452 hCoV-19/England/QEUH-B31AB2/2020\|EPI_ISL_646306\|20 | ···················· | ···················· | ····················· | ···················· | ···················· | ···················· |
| R | 3453 hCoV-19/England/ALDP-B23BB0/2020\|EPI_ISL_646307\|20 | ···················· | ···················· | ····················· | ···················· | ···················· | ···················· |
| R | 3454 hCoV-19/England/QEUH-B2DE71/2020\|EPI_ISL_646308\|20 | ···················· | ···················· | ····················· | ···················· | ···················· | ···················· |
| R | 3455 hCoV-19/England/ALDP-B23F2D/2020\|EPI_ISL_646309\|20 | ···················· | ···················· | ····················· | ···················· | ···················· | ···················· |
| R | 3456 hCoV-19/England/QEUH-B2E256/2020\|EPI_ISL_646310\|20 | ···················· | ···················· | ····················· | ···················· | ···················· | ···················· |
| R | 3457 hCoV-19/England/QEUH-B31B19/2020\|EPI_ISL_646311\|20 | ···················· | ···················· | ····················· | ···················· | ···················· | ···················· |
| R | 3458 hCoV-19/England/QEUH-B2F7FC/2020\|EPI_ISL_646312\|20 | ···················· | ···················· | ····················· | ···················· | ···················· | ···················· |
| R | 3459 hCoV-19/England/ALDP-B23C26/2020\|EPI_ISL_646313\|20 | ···················· | ···················· | ····················· | ···················· | ···················· | ···················· |
| R | 3460 hCoV-19/England/QEUH-B321AD/2020\|EPI_ISL_646315\|20 | ···················· | ···················· | ····················· | ···················· | ···················· | ···················· |
| R | 3461 hCoV-19/England/QEUH-B2E405/2020\|EPI_ISL_646316\|20 | ···················· | ···················· | ····················· | ···················· | ······T············· | ···················· |
| R | 3462 hCoV-19/England/QEUH-B31D6E/2020\|EPI_ISL_646317\|20 | ···················· | ···················· | ····················· | ···················· | ···················· | ···················· |
| R | 3463 hCoV-19/England/ALDP-B23B47/2020\|EPI_ISL_646318\|20 | ···················· | ···················· | ····················· | ···················· | ···················· | ···················· |
| R | 3464 hCoV-19/England/QEUH-B31B73/2020\|EPI_ISL_646319\|20 | ···················· | ···················· | ····················· | ···················· | ···················· | ···················· |
| R | 3465 hCoV-19/England/ALDP-B23B83/2020\|EPI_ISL_646320\|20 | ···················· | ···················· | ····················· | ···················· | ···················· | ···················· |
| R | 3466 hCoV-19/England/QEUH-B2E441/2020\|EPI_ISL_646321\|20 | ···················· | ···················· | ····················· | ···················· | ···················· | ···················· |
| R | 3467 hCoV-19/England/QEUH-B2E53F/2020\|EPI_ISL_646322\|20 | ···················· | ···················· | ····················· | ···················· | ···················· | ···················· |
| R | 3468 hCoV-19/England/QEUH-B32161/2020\|EPI_ISL_646324\|20 | ···················· | ···················· | ····················· | ···················· | ···················· | ···················· |
| R | 3469 hCoV-19/England/QEUH-B32134/2020\|EPI_ISL_646325\|20 | ···················· | ···················· | ····················· | ···················· | ···················· | ···················· |
| R | 3470 hCoV-19/England/QEUH-B31C25/2020\|EPI_ISL_646326\|20 | ···················· | ···················· | ····················· | ···················· | ···················· | ···················· |
| R | 3471 hCoV-19/England/ALDP-B23F78/2020\|EPI_ISL_646327\|20 | ···················· | ·A·················· | ····················· | ···················· | ···················· | ···················· |
| R | 3472 hCoV-19/England/QEUH-B318E5/2020\|EPI_ISL_646328\|20 | ···················· | ·A·················· | ····················· | ···················· | ···················· | ···A················ |
| R | 3473 hCoV-19/England/QEUH-B31180/2020\|EPI_ISL_646329\|20 | ···················· | ···················· | ····················· | ···················· | ···················· | ···················· |
| R | 3474 hCoV-19/England/QEUH-B2E432/2020\|EPI_ISL_646330\|20 | ···················· | ···················· | ····················· | ···················· | ···················· | ···················· |
| R | 3475 hCoV-19/England/QEUH-B31CF8/2020\|EPI_ISL_646331\|20 | ···················· | ···················· | ····················· | ···················· | ···················· | ···················· |
| R | 3476 hCoV-19/England/QEUH-B2F783/2020\|EPI_ISL_646332\|20 | ···················· | ···················· | ····················· | ···················· | ···················· | ···················· |
| R | 3477 hCoV-19/England/QEUH-B32082/2020\|EPI_ISL_646333\|20 | ···················· | ···················· | ····················· | ···················· | ···················· | ···················· |
| R | 3478 hCoV-19/England/ALDP-B23B38/2020\|EPI_ISL_646334\|20 | ···················· | ···················· | ····················· | ···················· | ···················· | ···················· |
| R | 3479 hCoV-19/England/QEUH-B31A3A/2020\|EPI_ISL_646335\|20 | ···················· | ·A·················· | ····················· | ···················· | ···················· | ···················· |
| R | 3480 hCoV-19/England/QEUH-B311AE/2020\|EPI_ISL_646336\|20 | ···················· | ···················· | ····················· | ···················· | ···················· | ···················· |
| R | 3481 hCoV-19/England/QEUH-B31A76/2020\|EPI_ISL_646337\|20 | ···················· | ···················· | ····················· | ···················· | ···················· | ···················· |
| R | 3482 hCoV-19/England/QEUH-B31BFB/2020\|EPI_ISL_646338\|20 | ···················· | ···················· | ····················· | ···················· | ···················· | ···················· |
| R | 3483 hCoV-19/England/QEUH-B32116/2020\|EPI_ISL_646339\|20 | ···················· | ···················· | ····················· | ···················· | ···················· | ···················· |
| R | 3484 hCoV-19/England/CAMC-B20F8A/2020\|EPI_ISL_647605\|20 | ···················· | ···················· | ····················· | ···················· | ···················· | ···················· |
| R | 3485 hCoV-19/England/ALDP-B1A958/2020\|EPI_ISL_647606\|20 | ···················· | ···················· | ····················· | ···················· | ···················· | ···················· |
| R | 3486 hCoV-19/England/CAMC-B21235/2020\|EPI_ISL_647607\|20 | ···················· | ·A·················· | ····················· | ···················· | ···················· | ···A················ |
| R | 3487 hCoV-19/England/ALDP-B1AA73/2020\|EPI_ISL_647608\|20 | ···················· | ···················· | ····················· | ···················· | ······T············· | ···················· |
| R | 3488 hCoV-19/England/CAMC-B21314/2020\|EPI_ISL_647609\|20 | ···················· | ···················· | ····················· | ···················· | ···················· | ···················· |
| R | 3489 hCoV-19/England/CAMC-B1FD2A/2020\|EPI_ISL_647614\|20 | ···················· | ···················· | ····················· | ···················· | ···················· | ···················· |
| R | 3490 hCoV-19/England/ALDP-B1A536/2020\|EPI_ISL_647615\|20 | ···················· | ···················· | ····················· | ···················· | ···················· | ···················· |
| R | 3491 hCoV-19/England/CAMC-B21183/2020\|EPI_ISL_647627\|20 | ···················· | ···················· | ····················· | ···················· | ···················· | ···················· |
| R | 3492 hCoV-19/England/CAMC-B1FC96/2020\|EPI_ISL_647628\|20 | ···················· | ···················· | ····················· | ···················· | ···················· | ···················· |
| R | 3493 hCoV-19/England/ALDP-B1A93A/2020\|EPI_ISL_647630\|20 | ···················· | ···················· | ····················· | ···················· | ···················· | ···················· |
| R | 3494 hCoV-19/England/CAMC-B205E2/2020\|EPI_ISL_647631\|20 | ···················· | ···················· | ····················· | ···················· | ···················· | ···················· |
| R | 3495 hCoV-19/England/ALDP-B1A5CD/2020\|EPI_ISL_647632\|20 | ···················· | ···················· | ····················· | ···················· | ···················· | ···················· |
| R | 3496 hCoV-19/England/CAMC-B210B3/2020\|EPI_ISL_647633\|20 | ···················· | ···················· | ····················· | ···················· | ···················· | ···················· |
| R | 3497 hCoV-19/England/ALDP-B1A509/2020\|EPI_ISL_647634\|20 | ···················· | ···················· | ····················· | ···················· | ···················· | ···················· |
| R | 3498 hCoV-19/England/ALDP-B1AACE/2020\|EPI_ISL_647635\|20 | ···················· | ···················· | ····················· | ···················· | ···················· | ···················· |
| R | 3499 hCoV-19/England/CAMC-B2129F/2020\|EPI_ISL_647636\|20 | ···················· | ···················· | ····················· | ···················· | ···················· | ···················· |
| R | 3500 hCoV-19/England/CAMC-B1FF24/2020\|EPI_ISL_647637\|20 | ···················· | ···················· | ····················· | ···················· | ···················· | ···················· |
| R | 3501 hCoV-19/England/ALDP-B1A85B/2020\|EPI_ISL_647638\|20 | ···················· | ···················· | ····················· | ···················· | ···················· | ···················· |
| R | 3502 hCoV-19/England/CAMC-B209AA/2020\|EPI_ISL_647639\|20 | ···················· | ···················· | ····················· | ···················· | ···················· | ···················· |
| R | 3503 hCoV-19/England/CAMC-B1FD66/2020\|EPI_ISL_647640\|20 | ···················· | ···················· | ····················· | ···················· | ···················· | ···················· |
| R | 3504 hCoV-19/England/CAMC-B20F5D/2020\|EPI_ISL_647642\|20 | ···················· | ···················· | ····················· | ···················· | ···················· | ···················· |
| R | 3505 hCoV-19/England/ALDP-B1AB34/2020\|EPI_ISL_647643\|20 | ···················· | ···················· | ····················· | ···················· | ······T············· | ···················· |
| R | 3506 hCoV-19/England/CAMC-B1FEBE/2020\|EPI_ISL_647644\|20 | ···················· | ···················· | ····················· | ···················· | ···················· | ···················· |
| R | 3507 hCoV-19/England/CAMC-B20F3F/2020\|EPI_ISL_647645\|20 | ···················· | ···················· | ····················· | ···················· | ···················· | ···················· |
| R | 3508 hCoV-19/England/CAMC-B1FF42/2020\|EPI_ISL_647646\|20 | ···················· | ···················· | ····················· | ···················· | ···················· | ···················· |
| R | 3509 hCoV-19/England/CAMC-B20B68/2020\|EPI_ISL_647647\|20 | ···················· | ·A·················· | ····················· | ···················· | ···················· | ···A················ |
| R | 3510 hCoV-19/England/CAMC-B20728/2020\|EPI_ISL_647648\|20 | ···················· | ···················· | ····················· | ···················· | ···················· | ···················· |
| R | 3511 hCoV-19/England/CAMC-B20904/2020\|EPI_ISL_647649\|20 | ···················· | ···················· | ····················· | ···················· | ···················· | ···················· |
| R | 3512 hCoV-19/England/ALDP-B1AA28/2020\|EPI_ISL_647650\|20 | ···················· | ···················· | ····················· | ···················· | ···················· | ···················· |
| R | 3513 hCoV-19/England/CAMC-B21569/2020\|EPI_ISL_647651\|20 | ···················· | ···················· | ····················· | ···················· | ···················· | ···················· |
| R | 3514 hCoV-19/England/CAMC-B21271/2020\|EPI_ISL_647652\|20 | ···················· | ···················· | ····················· | ···················· | ···················· | ···················· |
| R | 3515 hCoV-19/England/CAMC-B1FD93/2020\|EPI_ISL_647653\|20 | ···················· | ···················· | ····················· | ···················· | ···················· | ···················· |
| R | 3516 hCoV-19/England/CAMC-B1FECD/2020\|EPI_ISL_647654\|20 | ···················· | ···················· | ····················· | ···················· | ···················· | ···················· |
| R | 3517 hCoV-19/England/ALDP-B1A42A/2020\|EPI_ISL_647655\|20 | ···················· | ···················· | ····················· | ···················· | ···················· | ···················· |
| R | 3518 hCoV-19/England/CAMC-B212F9/2020\|EPI_ISL_647656\|20 | ···················· | ···················· | ····················· | ···················· | ···················· | ···················· |
| R | 3519 hCoV-19/England/CAMC-B1FCA5/2020\|EPI_ISL_647657\|20 | ···················· | ···················· | ····················· | ···················· | ···················· | ···················· |
| R | 3520 hCoV-19/England/CAMC-B1FC2D/2020\|EPI_ISL_647658\|20 | ···················· | ···················· | ····················· | ···················· | ···················· | ···················· |
| R | 3521 hCoV-19/England/ALDP-B1A7C7/2020\|EPI_ISL_647659\|20 | ···················· | ···················· | ····················· | ···················· | ···················· | ···················· |
| R | 3522 hCoV-19/England/CAMC-B20A01/2020\|EPI_ISL_647660\|20 | ···················· | ···················· | ····················· | ···················· | ···················· | ···················· |
| R | 3523 hCoV-19/England/CAMC-B20DAE/2020\|EPI_ISL_647661\|20 | ···················· | ···················· | ····················· | ···················· | ···················· | ···················· |
| R | 3524 hCoV-19/England/CAMC-B20913/2020\|EPI_ISL_647664\|20 | ···················· | ···················· | ····················· | ···················· | ···················· | ···················· |
| R | 3525 hCoV-19/England/CAMC-B20BFF/2020\|EPI_ISL_647665\|20 | ···················· | ···················· | ····················· | ···················· | ···················· | ···················· |
| R | 3526 hCoV-19/England/ALDP-B1A86A/2020\|EPI_ISL_647667\|20 | ···················· | ···················· | ····················· | ···················· | ···················· | ···················· |
| R | 3527 hCoV-19/England/ALDP-B1A4DF/2020\|EPI_ISL_647668\|20 | ···················· | ···················· | ····················· | ···················· | ···················· | ···················· |
| R | 3528 hCoV-19/England/CAMC-B2111A/2020\|EPI_ISL_647669\|20 | ···················· | ···················· | ····················· | ···················· | ···················· | ···················· |
| R | 3529 hCoV-19/USA/CA-ALSR-4230/2020\|EPI_ISL_648900\|2020- | ···················· | ·A·················· | ····················· | ···················· | ···················· | ···················· |
| R | 3530 hCoV-19/USA/CA-ALSR-4233/2020\|EPI_ISL_648901\|2020- | ···················· | ·A·················· | ····················· | ···················· | ···················· | ···················· |

| | | | | | | | |
|---|---|---|---|---|---|---|---|
| R | 3531 hCoV-19/USA/CA-ALSR-4235/2020\|EPI_ISL_648902\|2020- | ···················· | ·A·················· | ····················· | ···················· | ···················· | ···················· |
| R | 3532 hCoV-19/USA/CA-ALSR-4237/2020\|EPI_ISL_648904\|2020- | ···················· | ···················· | ····················· | ···················· | ···················· | ···················· |
| R | 3533 hCoV-19/USA/CA-ALSR-4238/2020\|EPI_ISL_648905\|2020- | ···················· | ···················· | ····················· | ···················· | ···················· | ···················· |
| R | 3534 hCoV-19/USA/CA-ALSR-4242/2020\|EPI_ISL_648906\|2020- | ···················· | ·A·················· | ····················· | ···················· | ···················· | ···················· |
| R | 3535 hCoV-19/USA/CA-ALSR-4243/2020\|EPI_ISL_648907\|2020- | ···················· | ·A·················· | ····················· | ···················· | ···················· | ···················· |
| R | 3536 hCoV-19/USA/CA-ALSR-4245/2020\|EPI_ISL_648908\|2020- | ·T·················· | ···················· | ····················· | ···················· | ···················· | ···················· |
| R | 3537 hCoV-19/USA/CA-ALSR-4247/2020\|EPI_ISL_648909\|2020- | ···················· | ·A·················· | ····················· | ···················· | ···················· | ···················· |
| R | 3538 hCoV-19/USA/CA-ALSR-4248/2020\|EPI_ISL_648910\|2020- | ···················· | ·A·················· | ····················· | ···················· | ···················· | ···················· |
| R | 3539 hCoV-19/USA/CA-ALSR-4249/2020\|EPI_ISL_648911\|2020- | ···················· | ·A·················· | ····················· | ···················· | ···················· | ···················· |
| R | 3540 hCoV-19/USA/CA-ALSR-4250/2020\|EPI_ISL_648912\|2020- | ···················· | ·A·················· | ····················· | ···················· | ···················· | ···················· |
| R | 3541 hCoV-19/USA/CA-ALSR-4251/2020\|EPI_ISL_648913\|2020- | ···················· | ···················· | ····················· | ···················· | ···················· | ···················· |
| R | 3542 hCoV-19/USA/CA-ALSR-4284/2020\|EPI_ISL_648936\|2020- | ···················· | ···················· | ····················· | ···················· | ···················· | ···················· |
| R | 3543 hCoV-19/USA/CA-ALSR-4285/2020\|EPI_ISL_648937\|2020- | ···················· | ·A·················· | ····················· | ···················· | ···················· | ···················· |
| R | 3544 hCoV-19/USA/CA-ALSR-4286/2020\|EPI_ISL_648938\|2020- | ···················· | ·A·················· | ····················· | ···················· | ···················· | ···················· |
| R | 3545 hCoV-19/USA/CA-ALSR-4287/2020\|EPI_ISL_648939\|2020- | ···················· | ···················· | ····················· | ···················· | ···················· | ···················· |
| R | 3546 hCoV-19/USA/CA-ALSR-4294/2020\|EPI_ISL_648946\|2020- | ···················· | ·A·················· | ····················· | ···················· | ···················· | ···················· |
| R | 3547 hCoV-19/USA/CA-ALSR-4308/2020\|EPI_ISL_648958\|2020- | ···················· | ·A·················· | ····················· | ···················· | ···················· | ···················· |
| R | 3548 hCoV-19/USA/CA-ALSR-4309/2020\|EPI_ISL_648959\|2020- | ···················· | ·A·················· | ····················· | ···················· | ···················· | ···················· |
| R | 3549 hCoV-19/USA/CA-ALSR-4310/2020\|EPI_ISL_648960\|2020- | ···················· | ·A·················· | ····················· | ···················· | ···················· | ···················· |
| R | 3550 hCoV-19/USA/CA-ALSR-4311/2020\|EPI_ISL_648961\|2020- | ···················· | ···················· | ····················· | ···················· | ···················· | ···················· |
| R | 3551 hCoV-19/USA/CA-ALSR-4313/2020\|EPI_ISL_648962\|2020- | ···················· | ·A·················· | ····················· | ···················· | ···················· | ···················· |
| R | 3552 hCoV-19/USA/CA-ALSR-4314/2020\|EPI_ISL_648963\|2020- | ···················· | ···················· | ····················· | ···················· | ···················· | ···················· |
| R | 3553 hCoV-19/USA/CA-ALSR-4315/2020\|EPI_ISL_648964\|2020- | ···················· | ·A·················· | ····················· | ···················· | ···················· | ···················· |
| R | 3554 hCoV-19/USA/CA-ALSR-4316/2020\|EPI_ISL_648965\|2020- | ···················· | ·A·················· | ····················· | ···················· | ···················· | ···················· |
| R | 3555 hCoV-19/USA/CA-ALSR-4317/2020\|EPI_ISL_648966\|2020- | ···················· | ···················· | ····················· | ···················· | ···················· | ···················· |
| R | 3556 hCoV-19/USA/CA-ALSR-4318/2020\|EPI_ISL_648967\|2020- | ···················· | ·A·················· | ····················· | ···················· | ···················· | ···················· |
| R | 3557 hCoV-19/USA/CA-ALSR-4319/2020\|EPI_ISL_648968\|2020- | ···················· | ···················· | ····················· | ···················· | ···················· | ···················· |
| R | 3558 hCoV-19/USA/CA-ALSR-4320/2020\|EPI_ISL_648969\|2020- | ···················· | ···················· | ····················· | ···················· | ···················· | ···················· |
| R | 3559 hCoV-19/USA/CA-ALSR-4322/2020\|EPI_ISL_648970\|2020- | ···················· | ·A·················· | ····················· | ···················· | ···················· | ···················· |
| R | 3560 hCoV-19/USA/CA-ALSR-4325/2020\|EPI_ISL_648971\|2020- | ···················· | ·A·················· | ····················· | ···················· | ···················· | ···················· |
| R | 3561 hCoV-19/USA/CA-ALSR-4326/2020\|EPI_ISL_648972\|2020- | ···················· | ·A·················· | ····················· | ···················· | ···················· | ···················· |
| R | 3562 hCoV-19/USA/CA-ALSR-4329/2020\|EPI_ISL_648974\|2020- | ···················· | ·A·················· | ····················· | ···················· | ···················· | ···················· |
| R | 3563 hCoV-19/USA/CA-ALSR-4331/2020\|EPI_ISL_648976\|2020- | ···················· | ·A·················· | ····················· | ···················· | ···················· | ···················· |
| R | 3564 hCoV-19/USA/CA-ALSR-4336/2020\|EPI_ISL_648978\|2020- | ···················· | ·A·················· | ····················· | ···················· | ···················· | ···················· |
| R | 3565 hCoV-19/USA/CA-ALSR-4337/2020\|EPI_ISL_648979\|2020- | ···················· | ·A·················· | ····················· | ···················· | ···················· | ···················· |
| R | 3566 hCoV-19/USA/CA-ALSR-4340/2020\|EPI_ISL_648980\|2020- | ···················· | ···················· | ····················· | ···················· | ···················· | ···················· |
| R | 3567 hCoV-19/USA/CA-ALSR-4342/2020\|EPI_ISL_648981\|2020- | ···················· | ·A·················· | ····················· | ···················· | ···················· | ···················· |
| R | 3568 hCoV-19/USA/CA-ALSR-4343/2020\|EPI_ISL_648982\|2020- | ···················· | ·A·················· | ····················· | ···················· | ···················· | ···················· |
| R | 3569 hCoV-19/USA/CA-ALSR-4344/2020\|EPI_ISL_648983\|2020- | ···················· | ·A·················· | ····················· | ···················· | ···················· | ···················· |
| R | 3570 hCoV-19/USA/CA-ALSR-4347/2020\|EPI_ISL_648985\|2020- | ···················· | ·A·················· | ····················· | ···················· | ···················· | ···················· |
| R | 3571 hCoV-19/USA/CA-ALSR-4351/2020\|EPI_ISL_648986\|2020- | ···················· | ·A·················· | ····················· | ···················· | ···················· | ···················· |
| R | 3572 hCoV-19/USA/CA-ALSR-4356/2020\|EPI_ISL_648987\|2020- | ···················· | ···················· | ····················· | ···················· | ···················· | ···················· |
| R | 3573 hCoV-19/USA/CA-ALSR-4358/2020\|EPI_ISL_648988\|2020- | ···················· | ·A·················· | ····················· | ···················· | ···················· | ···················· |
| R | 3574 hCoV-19/USA/CA-ALSR-4359/2020\|EPI_ISL_648989\|2020- | ···················· | ·A·················· | ····················· | ···················· | ···················· | ···················· |
| R | 3575 hCoV-19/USA/CA-ALSR-4364/2020\|EPI_ISL_648990\|2020- | ···················· | ·A·················· | ····················· | ···················· | ···················· | ···················· |
| R | 3576 hCoV-19/USA/CA-ALSR-4365/2020\|EPI_ISL_648991\|2020- | ···················· | ·A·················· | ····················· | ···················· | ···················· | ···················· |
| R | 3577 hCoV-19/USA/CA-ALSR-4366/2020\|EPI_ISL_648992\|2020- | ···················· | ·A·················· | ····················· | ···················· | ···················· | ···················· |
| R | 3578 hCoV-19/USA/CA-ALSR-4369/2020\|EPI_ISL_648993\|2020- | ···················· | ·A·················· | ····················· | ···················· | ···················· | ···················· |
| R | 3579 hCoV-19/USA/CA-ALSR-4374/2020\|EPI_ISL_648994\|2020- | ···················· | ·A·················· | ····················· | ···················· | ···················· | ···················· |
| R | 3580 hCoV-19/USA/CA-ALSR-4376/2020\|EPI_ISL_648995\|2020- | ···················· | ·A·················· | ····················· | ···················· | ···················· | ···················· |
| R | 3581 hCoV-19/USA/CA-ALSR-4380/2020\|EPI_ISL_648996\|2020- | ···················· | ·A·················· | ····················· | ···················· | ···················· | ···················· |
| R | 3582 hCoV-19/USA/CA-ALSR-4381/2020\|EPI_ISL_648997\|2020- | ···················· | ·A·················· | ····················· | ···················· | ···················· | ···················· |
| R | 3583 hCoV-19/USA/CA-ALSR-4384/2020\|EPI_ISL_648998\|2020- | ···················· | ·A·················· | ····················· | ···················· | ···················· | ···················· |
| R | 3584 hCoV-19/USA/CA-ALSR-4385/2020\|EPI_ISL_648999\|2020- | ···················· | ·A·················· | ····················· | ···················· | ···················· | ···················· |
| R | 3585 hCoV-19/South Africa/NHLS-UCT-GP-5227/2020\|EPI_ISL | ···················· | ···················· | ····················· | ···················· | ·NN················· | ···················· |
| R | 3586 hCoV-19/South Africa/NHLS-UCT-GS-1770/2020\|EPI_ISL | ···················· | ·A·················· | ····················· | ···················· | ···················· | ···················· |
| R | 3587 hCoV-19/South Africa/NHLS-UCT-GS-1772/2020\|EPI_ISL | ···················· | ·A·················· | ····················· | ···················· | ···················· | ···················· |
| R | 3588 hCoV-19/South Africa/NHLS-UCT-GS-1777/2020\|EPI_ISL | ···················· | ···················· | ····················· | ···················· | ···················· | ···················· |
| R | 3589 hCoV-19/South Africa/NHLS-UCT-GS-1779/2020\|EPI_ISL | ···················· | ···················· | ····················· | ···················· | ···················· | ···················· |
| R | 3590 hCoV-19/South Africa/NHLS-UCT-GS-1780/2020\|EPI_ISL | ···················· | ···················· | ····················· | ···················· | ···················· | ···················· |
| R | 3591 hCoV-19/mink/Denmark/mDK-21/2020\|EPI_ISL_641414\|20 | ···················· | ···················· | ····················· | ···················· | ···················· | ···················· |
| R | 3595 hCoV-19/mink/Denmark/mDK-29/2020\|EPI_ISL_641422\|20 | ···················· | ···················· | ····················· | ···················· | ···················· | ···················· |
| R | 3597 hCoV-19/England/QEUH-B12510/2020\|EPI_ISL_642710\|20 | ···················· | ···················· | ····················· | ···················· | ···················· | ···················· |
| R | 3598 hCoV-19/England/QEUH-B0F17C/2020\|EPI_ISL_642711\|20 | ···················· | ···················· | ····················· | ···················· | ···················· | ···················· |
| R | 3599 hCoV-19/England/QEUH-B12BA5/2020\|EPI_ISL_642712\|20 | ···················· | ···················· | ····················· | ···················· | ···················· | ···················· |
| R | 3600 hCoV-19/England/QEUH-B0F22E/2020\|EPI_ISL_642713\|20 | ···················· | ···················· | ····················· | ···················· | ···················· | ···················· |
| R | 3601 hCoV-19/England/QEUH-B1213A/2020\|EPI_ISL_642714\|20 | ···················· | ···················· | ····················· | ···················· | ···················· | ···················· |
| R | 3602 hCoV-19/England/QEUH-B1248C/2020\|EPI_ISL_642715\|20 | ···················· | ·A·················· | ····················· | ···················· | ···················· | ···A················ |
| R | 3603 hCoV-19/England/QEUH-B112FC/2020\|EPI_ISL_642716\|20 | ···················· | ···················· | ····················· | ···················· | ···················· | ···················· |
| R | 3604 hCoV-19/England/QEUH-B123E9/2020\|EPI_ISL_642717\|20 | ···················· | ···················· | ····················· | ···················· | ···················· | ···················· |
| R | 3605 hCoV-19/England/QEUH-B1439C/2020\|EPI_ISL_642718\|20 | ···················· | ···················· | ····················· | ···················· | ···················· | ···················· |
| R | 3606 hCoV-19/England/QEUH-B10A5F/2020\|EPI_ISL_642719\|20 | ···················· | ···················· | ····················· | ···················· | ···················· | ···················· |
| R | 3607 hCoV-19/England/QEUH-B100A8/2020\|EPI_ISL_642720\|20 | ···················· | ···················· | ····················· | ···················· | ···················· | ···················· |
| R | 3608 hCoV-19/England/QEUH-B112B0/2020\|EPI_ISL_642721\|20 | ···················· | ···················· | ····················· | ···················· | ···················· | ···················· |
| R | 3609 hCoV-19/England/QEUH-B1438D/2020\|EPI_ISL_642722\|20 | ···················· | ···················· | ····················· | ···················· | ···················· | ···················· |
| R | 3610 hCoV-19/England/QEUH-B0EE3A/2020\|EPI_ISL_642723\|20 | ···················· | ···················· | ····················· | ···················· | ···················· | ···················· |
| R | 3611 hCoV-19/England/QEUH-B12000/2020\|EPI_ISL_642724\|20 | ···················· | ···················· | ····················· | ···················· | ···················· | ···················· |
| R | 3612 hCoV-19/England/QEUH-B100E4/2020\|EPI_ISL_642725\|20 | ···················· | ···················· | ····················· | ···················· | ···················· | ···················· |
| R | 3613 hCoV-19/England/QEUH-B12352/2020\|EPI_ISL_642726\|20 | ···················· | ···················· | ····················· | ···················· | ···················· | ···················· |
| R | 3614 hCoV-19/England/QEUH-B1205B/2020\|EPI_ISL_642727\|20 | ···················· | ···················· | ····················· | ···················· | ···················· | ···················· |

```
R 3615 hCoV-19/England/QEUH-B12A3F/2020|EPI_ISL_642729|20 ···················· ···················· ····················· ···················· ···················· ····················
R 3616 hCoV-19/England/QEUH-B144F3/2020|EPI_ISL_642730|20 ···················· ···················· ····················· ···················· ···················· ····················
R 3617 hCoV-19/England/QEUH-B10BF2/2020|EPI_ISL_642731|20 ···················· ···················· ····················· ···················· ···················· ····················
R 3618 hCoV-19/England/QEUH-B100C6/2020|EPI_ISL_642732|20 ···················· ···················· ····················· ···················· ···················· ····················
R 3619 hCoV-19/England/QEUH-B12ED9/2020|EPI_ISL_642733|20 ···················· ···················· ····················· ···················· ···················· ····················
R 3620 hCoV-19/England/QEUH-B10A04/2020|EPI_ISL_642735|20 ···················· ···················· ····················· ···················· ···················· ····················
R 3621 hCoV-19/England/QEUH-B125E3/2020|EPI_ISL_642736|20 ···················· ···················· ····················· ···················· ···················· ····················
R 3622 hCoV-19/England/QEUH-B1455A/2020|EPI_ISL_642737|20 ···················· ···················· ····················· ···················· ···················· ····················
R 3623 hCoV-19/England/QEUH-B0F23D/2020|EPI_ISL_642738|20 ···················· ···················· ····················· ···················· ···················· ····················
R 3624 hCoV-19/England/QEUH-B12E9D/2020|EPI_ISL_642739|20 ·····G·············· ···················· ····················· ···················· ···················· ····················
R 3625 hCoV-19/England/QEUH-B0F200/2020|EPI_ISL_642740|20 ···················· ···················· ····················· ···················· ···················· ····················
R 3626 hCoV-19/England/QEUH-B123CB/2020|EPI_ISL_642741|20 ···················· ···················· ····················· ···················· ···················· ····················
R 3627 hCoV-19/England/QEUH-B1011E/2020|EPI_ISL_642742|20 ·····G·············· ···················· ····················· ···················· ···················· ····················
R 3628 hCoV-19/England/QEUH-B143E7/2020|EPI_ISL_642743|20 ···················· ···················· ····················· ···················· ···················· ····················
R 3629 hCoV-19/England/QEUH-B124E6/2020|EPI_ISL_642744|20 ···················· ···················· ····················· ···················· ···················· ····················
R 3630 hCoV-19/England/QEUH-B10C1D/2020|EPI_ISL_642745|20 ···················· ···················· ····················· ···················· ···················· ····················
R 3631 hCoV-19/England/QEUH-B10A31/2020|EPI_ISL_642746|20 ···················· ···················· ····················· ···················· ···················· ····················
R 3632 hCoV-19/England/QEUH-B126C2/2020|EPI_ISL_642747|20 ···················· ···················· ····················· ···················· ···················· ····················
R 3633 hCoV-19/England/QEUH-B12097/2020|EPI_ISL_642748|20 ···················· ···················· ····················· ···················· ···················· ····················
R 3634 hCoV-19/England/QEUH-B12DCD/2020|EPI_ISL_642749|20 ···················· ···················· ····················· ···················· ···················· ····················
R 3635 hCoV-19/England/QEUH-B12BB4/2020|EPI_ISL_642750|20 ···················· ···················· ····················· ···················· ···················· ····················
R 3636 hCoV-19/England/QEUH-B11344/2020|EPI_ISL_642752|20 ···················· ···················· ····················· ···················· ···················· ····················
R 3637 hCoV-19/England/QEUH-B12DBE/2020|EPI_ISL_642753|20 ···················· ···················· ····················· ···················· ···················· ····················
R 3638 hCoV-19/England/QEUH-B10B6B/2020|EPI_ISL_642754|20 ···················· ···················· ····················· ···················· ···················· ····················
R 3639 hCoV-19/England/QEUH-B1452D/2020|EPI_ISL_642755|20 ···················· ···················· ····················· ···················· ···················· ····················
R 3640 hCoV-19/England/QEUH-B12A11/2020|EPI_ISL_642756|20 ···················· ···················· ····················· ···················· ···················· ····················
R 3641 hCoV-19/England/QEUH-B10C3B/2020|EPI_ISL_642757|20 ···················· ···················· ····················· ···················· ···················· ····················
R 3642 hCoV-19/England/QEUH-B1448A/2020|EPI_ISL_642758|20 ···················· ···················· ····················· ···················· ···················· ····················
R 3643 hCoV-19/England/QEUH-B12EF7/2020|EPI_ISL_642759|20 ···················· ···················· ····················· ···················· ···················· ····················
R 3644 hCoV-19/England/QEUH-B1447B/2020|EPI_ISL_642760|20 ···················· ···················· ····················· ···················· ···················· ····················
R 3645 hCoV-19/England/MILK-B02A6F/2020|EPI_ISL_642762|20 ···················· ···················· ····················· ···················· ······T············· ····················
R 3646 hCoV-19/England/MILK-B02A41/2020|EPI_ISL_642763|20 ···················· ···················· ····················· ···················· ···················· ····················
R 3647 hCoV-19/England/MILK-B022EF/2020|EPI_ISL_642764|20 ···················· ···················· ····················· ···················· ···················· ····················
R 3648 hCoV-19/England/MILK-B0220D/2020|EPI_ISL_642765|20 ···················· ···················· ····················· ···················· ···················· ····················
R 3649 hCoV-19/England/MILK-B0604B/2020|EPI_ISL_642766|20 ···················· ···················· ····················· ···················· ···················· ····················
R 3650 hCoV-19/England/MILK-B02294/2020|EPI_ISL_642767|20 ···················· ···················· ····················· ···················· ···················· ····················
R 3651 hCoV-19/England/MILK-B05EBB/2020|EPI_ISL_642768|20 ···················· ···················· ····················· ···················· ······T············· ····················
R 3652 hCoV-19/England/MILK-B02355/2020|EPI_ISL_642769|20 ···················· ···················· ····················· ···················· ···················· ····················
R 3653 hCoV-19/England/MILK-B02DB1/2020|EPI_ISL_642770|20 ···················· ···················· ····················· ···················· ···················· ····················
R 3654 hCoV-19/England/MILK-B0256E/2020|EPI_ISL_642771|20 ···················· ···················· ····················· ···················· ······T············· ····················
R 3655 hCoV-19/England/MILK-B02382/2020|EPI_ISL_642772|20 ···················· ···················· ····················· ···················· ···················· ····················
R 3656 hCoV-19/England/MILK-B0211F/2020|EPI_ISL_642773|20 ···················· ···················· ····················· ···················· ······T············· ····················
R 3657 hCoV-19/England/MILK-B02540/2020|EPI_ISL_642774|20 ···················· ···················· ····················· ···················· ···················· ····················
R 3658 hCoV-19/England/MILK-B01EEC/2020|EPI_ISL_642775|20 ···················· ·A·······A·········· ····················· ···················· ···················· ····················
R 3659 hCoV-19/England/QEUH-B31C9E/2020|EPI_ISL_646219|20 ···················· ···················· ····················· ···················· ···················· ····················
R 3660 hCoV-19/England/QEUH-B31AA3/2020|EPI_ISL_646220|20 ···················· ···················· ····················· ···················· ···················· ····················
R 3661 hCoV-19/England/QEUH-B32152/2020|EPI_ISL_646221|20 ···················· ···················· ····················· ···················· ···················· ····················
R 3662 hCoV-19/England/QEUH-B2E10E/2020|EPI_ISL_646222|20 ···················· ···················· ····················· ···················· ···················· ····················
R 3663 hCoV-19/England/ALDP-B24047/2020|EPI_ISL_646223|20 ···················· ······R············· ····················· ···················· ···················· ····················
R 3664 hCoV-19/England/ALDP-B2401A/2020|EPI_ISL_646224|20 ···················· ···················· ····················· ···················· ···················· ····················
R 3665 hCoV-19/England/QEUH-B2E46F/2020|EPI_ISL_646225|20 ···················· ···················· ····················· ···················· ···················· ····················
R 3666 hCoV-19/England/QEUH-B2E423/2020|EPI_ISL_646226|20 ···················· ···················· ····················· ···················· ···················· ····················
R 3667 hCoV-19/England/QEUH-B319E2/2020|EPI_ISL_646227|20 ···················· ···················· ····················· ···················· ···················· ····················
R 3668 hCoV-19/England/QEUH-B31E4D/2020|EPI_ISL_646228|20 ···················· ···················· ····················· ···················· ···················· ····················
R 3669 hCoV-19/England/QEUH-B31A58/2020|EPI_ISL_646229|20 ···················· ···················· ····················· ···················· ···················· ····················
R 3670 hCoV-19/England/QEUH-B31CE9/2020|EPI_ISL_646230|20 ···················· ···················· ····················· ···················· ···················· ····················
R 3671 hCoV-19/England/ALDP-B23BCF/2020|EPI_ISL_646231|20 ···················· ···················· ····················· ···················· ···················· ····················
R 3672 hCoV-19/England/QEUH-B31F3B/2020|EPI_ISL_646233|20 ···················· ···················· ····················· ···················· ···················· ····················
R 3673 hCoV-19/England/QEUH-B31DAA/2020|EPI_ISL_646234|20 ···················· ···················· ····················· ···················· ···················· ····················
R 3674 hCoV-19/England/ALDP-B23E99/2020|EPI_ISL_646235|20 ···················· ···················· ····················· ···················· ···················· ····················
R 3675 hCoV-19/England/QEUH-B31B0A/2020|EPI_ISL_646236|20 ···················· ·········A·········· ····················· ···················· ······T············· ····················
R 3676 hCoV-19/England/QEUH-B2E0B6/2020|EPI_ISL_646237|20 ···················· ···················· ····················· ···················· ···················· ····················
R 3677 hCoV-19/England/QEUH-B31AC1/2020|EPI_ISL_646238|20 ···················· ·A·················· ····················· ···················· ···················· ····················
R 3678 hCoV-19/England/QEUH-B31B46/2020|EPI_ISL_646239|20 ···················· ···················· ····················· ···················· ···················· ····················
R 3679 hCoV-19/England/QEUH-B31FA4/2020|EPI_ISL_646240|20 ···················· ···················· ····················· ···················· ···················· ····················
R 3680 hCoV-19/England/QEUH-B2E098/2020|EPI_ISL_646241|20 ···················· ···················· ····················· ···················· ···················· ····················
R 3681 hCoV-19/England/ALDP-B23B65/2020|EPI_ISL_646242|20 ···················· ···················· ····················· ···················· ···················· ····················
R 3682 hCoV-19/England/QEUH-B2E186/2020|EPI_ISL_646243|20 ···················· ···················· ····················· ···················· ···················· ····················
R 3683 hCoV-19/England/QEUH-B319C4/2020|EPI_ISL_646244|20 ···················· ···················· ····················· ···················· ···················· ····················
R 3684 hCoV-19/England/ALDP-B23D14/2020|EPI_ISL_646245|20 ···················· ···················· ····················· ···················· ···················· ····················
R 3685 hCoV-19/England/QEUH-B31A49/2020|EPI_ISL_646246|20 ···················· ···················· ····················· ···················· ···················· ····················
R 3686 hCoV-19/England/QEUH-B2E450/2020|EPI_ISL_646247|20 ···················· ···················· ····················· ···················· ···················· ····················
R 3687 hCoV-19/England/ALDP-B23C9F/2020|EPI_ISL_646248|20 ···················· ···················· ····················· ···················· ···················· ····················
R 3688 hCoV-19/England/ALDP-B23E5D/2020|EPI_ISL_646249|20 ···················· ···················· ····················· ···················· ···················· ····················
R 3689 hCoV-19/England/QEUH-B31092/2020|EPI_ISL_646250|20 ···················· ···················· ····················· ···················· ···················· ····················
R 3690 hCoV-19/England/ALDP-B23E7B/2020|EPI_ISL_646252|20 ···················· ···················· ····················· ···················· ···················· ····················
R 3691 hCoV-19/England/QEUH-B2E11D/2020|EPI_ISL_646253|20 ···················· ···················· ····················· ···················· ···················· ····················
R 3692 hCoV-19/England/QEUH-B2E03E/2020|EPI_ISL_646254|20 ···················· ···················· ····················· ···················· ···················· ····················
R 3693 hCoV-19/England/QEUH-B32125/2020|EPI_ISL_646255|20 ···················· ···················· ····················· ···················· ···················· ····················
R 3694 hCoV-19/England/QEUH-B320DD/2020|EPI_ISL_646256|20 ···················· ···················· ····················· ···················· ···················· ····················
```

| | | | | | | | |
|---|---|---|---|---|---|---|---|
| R | 3695 hCoV-19/England/QEUH-B32064/2020\|EPI_ISL_646257\|20 | ···················· | ···················· | ···················· | ···················· | ···················· | ···················· |
| R | 3696 hCoV-19/England/QEUH-B32019/2020\|EPI_ISL_646258\|20 | ···················· | ···················· | ···················· | ···················· | ·····T·············· | ···················· |
| R | 3697 hCoV-19/England/QEUH-B2F7ED/2020\|EPI_ISL_646259\|20 | ···················· | ···················· | ···················· | ···················· | ···················· | ···················· |
| R | 3698 hCoV-19/England/QEUH-B31D31/2020\|EPI_ISL_646260\|20 | ···················· | ···················· | ···················· | ···················· | ···················· | ···················· |
| R | 3699 hCoV-19/England/ALDP-B2400B/2020\|EPI_ISL_646261\|20 | ···················· | ···················· | ···················· | ···················· | ···················· | ···················· |
| R | 3700 hCoV-19/England/QEUH-B31BBF/2020\|EPI_ISL_646262\|20 | ···················· | ·A·················· | ···················· | ···················· | ···················· | ···················· |
| R | 3701 hCoV-19/England/QEUH-B31E7A/2020\|EPI_ISL_646263\|20 | ···················· | ·A·················· | ···················· | ···················· | ···················· | ···················· |
| R | 3702 hCoV-19/England/QEUH-B2E13B/2020\|EPI_ISL_646264\|20 | ···················· | ···················· | ···················· | ···················· | ···················· | ···················· |
| R | 3703 hCoV-19/England/QEUH-B31EA7/2020\|EPI_ISL_646265\|20 | ···················· | ···················· | ···················· | ···················· | ···················· | ···················· |
| R | 3704 hCoV-19/England/QEUH-B2DE62/2020\|EPI_ISL_646266\|20 | ···················· | ···················· | ···················· | ···················· | ···················· | ···················· |
| R | 3705 hCoV-19/England/QEUH-B318B8/2020\|EPI_ISL_646267\|20 | ···················· | ···················· | ···················· | ···················· | ···················· | ···················· |
| R | 3706 hCoV-19/England/ALDP-B23C08/2020\|EPI_ISL_646268\|20 | ···················· | ···················· | ···················· | ···················· | ···················· | ···················· |
| R | 3707 hCoV-19/England/ALDP-B23CF9/2020\|EPI_ISL_646269\|20 | ···················· | ···················· | ···················· | ···················· | ···················· | ···················· |
| R | 3708 hCoV-19/England/QEUH-B31EB6/2020\|EPI_ISL_646270\|20 | ···················· | ···················· | ···················· | ···················· | ···················· | ···················· |
| R | 3709 hCoV-19/England/ALDP-B23D8D/2020\|EPI_ISL_646271\|20 | ···················· | ···················· | ···················· | ···················· | ···················· | ···················· |
| R | 3710 hCoV-19/England/QEUH-B31126/2020\|EPI_ISL_646272\|20 | ···················· | ···················· | ···················· | ···················· | ···················· | ···················· |
| R | 3711 hCoV-19/England/QEUH-B31B28/2020\|EPI_ISL_646273\|20 | ···················· | ···················· | ···················· | ···················· | ···················· | ···················· |
| R | 3712 hCoV-19/England/QEUH-B32091/2020\|EPI_ISL_646274\|20 | ···················· | ···················· | ···················· | ···················· | ···················· | ···················· |
| R | 3713 hCoV-19/England/QEUH-B31BEC/2020\|EPI_ISL_646275\|20 | ···················· | ···················· | ···················· | ···················· | ···················· | ···················· |
| R | 3714 hCoV-19/England/ALDP-B23D05/2020\|EPI_ISL_646276\|20 | ···················· | ·A·················· | ···················· | ···················· | ···················· | ··A················· |
| R | 3715 hCoV-19/England/ALDP-B23C62/2020\|EPI_ISL_646277\|20 | ···················· | ···················· | ···················· | ···················· | ···················· | ···················· |
| R | 3716 hCoV-19/England/ALDP-B23FE1/2020\|EPI_ISL_646279\|20 | ···················· | ···················· | ···················· | ···················· | ···················· | ···················· |
| R | 3717 hCoV-19/England/QEUH-B320CE/2020\|EPI_ISL_646280\|20 | ···················· | ···················· | ···················· | ···················· | ···················· | ···················· |
| R | 3718 hCoV-19/England/QEUH-B31135/2020\|EPI_ISL_646281\|20 | ···················· | ···················· | ···················· | ···················· | ···················· | ···················· |
| R | 3719 hCoV-19/England/ALDP-B24074/2020\|EPI_ISL_646283\|20 | ···················· | ···················· | ···················· | ···················· | ···················· | ···················· |
| R | 3720 hCoV-19/England/QEUH-B2DDA1/2020\|EPI_ISL_646284\|20 | ···················· | ···················· | ···················· | ···················· | ···················· | ···················· |
| R | 3721 hCoV-19/England/QEUH-B31900/2020\|EPI_ISL_646285\|20 | ···················· | ·A·················· | ···················· | ···················· | ···················· | ··A················· |
| R | 3722 hCoV-19/England/QEUH-B2E238/2020\|EPI_ISL_646286\|20 | ···················· | ···················· | ···················· | ···················· | ···················· | ···················· |
| R | 3723 hCoV-19/England/QEUH-B31D8C/2020\|EPI_ISL_646287\|20 | ···················· | ···················· | ···················· | ···················· | ···················· | ···················· |
| R | 3724 hCoV-19/England/QEUH-B31C61/2020\|EPI_ISL_646288\|20 | ···················· | ···················· | ···················· | ···················· | ···················· | ···················· |
| R | 3725 hCoV-19/England/QEUH-B31BA0/2020\|EPI_ISL_646289\|20 | ···················· | ···················· | ···················· | ···················· | ···T················ | ···················· |
| R | 3726 hCoV-19/England/QEUH-B311CC/2020\|EPI_ISL_646290\|20 | ···················· | ···················· | ···················· | ···················· | ···················· | ···················· |
| R | 3727 hCoV-19/England/ALDP-B23CDB/2020\|EPI_ISL_646291\|20 | ···················· | ···················· | ···················· | ···················· | ···················· | ···················· |
| R | 3728 hCoV-19/England/QEUH-B2E159/2020\|EPI_ISL_646292\|20 | ···················· | ···················· | ···················· | ···················· | ···················· | ···················· |
| R | 3729 hCoV-19/England/QEUH-B32107/2020\|EPI_ISL_646293\|20 | ···················· | ···················· | ···················· | ···················· | ···················· | ···················· |
| R | 3730 hCoV-19/England/QEUH-B3192E/2020\|EPI_ISL_646294\|20 | ···················· | ···················· | ···················· | ···················· | ···················· | ···················· |
| R | 3731 hCoV-19/England/QEUH-B2E274/2020\|EPI_ISL_646295\|20 | ···················· | ···················· | ···················· | ···················· | ···················· | ···················· |
| R | 3732 hCoV-19/England/QEUH-B31DF5/2020\|EPI_ISL_646296\|20 | ···················· | ···················· | ···················· | ···················· | ···················· | ···················· |
| R | 3733 hCoV-19/England/ALDP-B23CEA/2020\|EPI_ISL_646297\|20 | ···················· | ···················· | ···················· | ···················· | ···················· | ···················· |
| R | 3734 hCoV-19/England/QEUH-B2DDDE/2020\|EPI_ISL_646298\|20 | ···················· | ···················· | ···················· | ···················· | ···················· | ···················· |
| R | 3735 hCoV-19/England/ALDP-B240FC/2020\|EPI_ISL_646299\|20 | ···················· | ···················· | ···················· | ···················· | ···················· | ···················· |
| R | 3736 hCoV-19/England/CAMC-B20263/2020\|EPI_ISL_647552\|20 | ···················· | ···················· | ···················· | ···················· | ···················· | ···················· |
| R | 3737 hCoV-19/England/CAMC-B20755/2020\|EPI_ISL_647554\|20 | ···················· | ···················· | ···················· | ···················· | ···················· | ···················· |
| R | 3738 hCoV-19/England/CAMC-B2154B/2020\|EPI_ISL_647555\|20 | ···················· | ···················· | ···················· | ···················· | ···················· | ···················· |
| R | 3739 hCoV-19/England/CAMC-B211FC/2020\|EPI_ISL_647557\|20 | ···················· | ···················· | ···················· | ···················· | ···················· | ···················· |
| R | 3740 hCoV-19/England/ALDP-B1A7B8/2020\|EPI_ISL_647558\|20 | ···················· | ···················· | ···················· | ···················· | ···················· | ···················· |
| R | 3741 hCoV-19/England/CAMC-B20CB0/2020\|EPI_ISL_647559\|20 | ···················· | ···················· | ···················· | ···················· | ···················· | ···················· |
| R | 3742 hCoV-19/England/CAMC-B1FF8E/2020\|EPI_ISL_647561\|20 | ···················· | ···················· | ···················· | ···················· | ···················· | ···················· |
| R | 3743 hCoV-19/England/CAMC-B20AC5/2020\|EPI_ISL_647563\|20 | ···················· | ···················· | ···················· | ···················· | ···················· | ···················· |
| R | 3744 hCoV-19/England/CAMC-B1FBA8/2020\|EPI_ISL_647564\|20 | ···················· | ···················· | ···················· | ···················· | ···················· | ···················· |
| R | 3745 hCoV-19/England/CAMC-B21578/2020\|EPI_ISL_647565\|20 | ···················· | ···················· | ···················· | ···················· | ···················· | ···················· |
| R | 3746 hCoV-19/England/CAMC-B20D35/2020\|EPI_ISL_647566\|20 | ···················· | ···················· | ···················· | ···················· | ···················· | ···················· |
| R | 3747 hCoV-19/England/CAMC-B20A98/2020\|EPI_ISL_647567\|20 | ···················· | ···················· | ···················· | ···················· | ···················· | ···················· |
| R | 3748 hCoV-19/England/CAMC-B1FB8A/2020\|EPI_ISL_647568\|20 | ···················· | ···················· | ···················· | ···················· | ·····T·············· | ···················· |
| R | 3749 hCoV-19/England/CAMC-B20834/2020\|EPI_ISL_647569\|20 | ···················· | ···················· | ···················· | ···················· | ·····T·············· | ···················· |
| R | 3750 hCoV-19/England/CAMC-B1FF7F/2020\|EPI_ISL_647570\|20 | ···················· | ···················· | ···················· | ···················· | ···················· | ···················· |
| R | 3751 hCoV-19/England/CAMC-B20746/2020\|EPI_ISL_647571\|20 | ···················· | ···················· | ···················· | ···················· | ···················· | ···················· |
| R | 3752 hCoV-19/England/CAMC-B2155A/2020\|EPI_ISL_647574\|20 | ···················· | ···················· | ···················· | ···················· | ···················· | ···················· |
| R | 3753 hCoV-19/England/CAMC-B202DC/2020\|EPI_ISL_647575\|20 | ···················· | ···················· | ···················· | ···················· | ·····T·············· | ···················· |
| R | 3754 hCoV-19/England/ALDP-B1A8E2/2020\|EPI_ISL_647576\|20 | ···················· | ···················· | ···················· | ···················· | ···················· | ···················· |
| R | 3755 hCoV-19/England/CAMC-B2000F/2020\|EPI_ISL_647577\|20 | ···················· | ···················· | ···················· | ···················· | ···················· | ···················· |
| R | 3756 hCoV-19/England/ALDP-B1A4EE/2020\|EPI_ISL_647579\|20 | ···················· | ···················· | ···················· | ···················· | ···················· | ···················· |
| R | 3757 hCoV-19/England/CAMC-B20DDB/2020\|EPI_ISL_647581\|20 | ···················· | ···················· | ···················· | ···················· | ···················· | ···················· |
| R | 3758 hCoV-19/England/CAMC-B20F20/2020\|EPI_ISL_647582\|20 | ···················· | ···················· | ···················· | ···················· | ···················· | ···················· |
| R | 3759 hCoV-19/England/CAMC-B21208/2020\|EPI_ISL_647583\|20 | ···················· | ···················· | ···················· | ···················· | ···················· | ···················· |
| R | 3760 hCoV-19/England/ALDP-B1A7A9/2020\|EPI_ISL_647584\|20 | ···················· | ···················· | ···················· | ···················· | ···················· | ···················· |
| R | 3761 hCoV-19/England/CAMC-B20922/2020\|EPI_ISL_647585\|20 | ···················· | ···················· | ···················· | ···················· | ···················· | ··A················· |
| R | 3762 hCoV-19/England/CAMC-B1FFD9/2020\|EPI_ISL_647586\|20 | ···················· | ···················· | ···················· | ···················· | ·····T·············· | ···················· |
| R | 3763 hCoV-19/England/CAMC-B1FBF3/2020\|EPI_ISL_647587\|20 | ···················· | ···················· | ···················· | ···················· | ·····T·············· | ···················· |
| R | 3764 hCoV-19/England/CAMC-B1FFBB/2020\|EPI_ISL_647588\|20 | ···················· | ·A·················· | ···················· | ···················· | ···················· | ··A················· |
| R | 3765 hCoV-19/England/CAMC-B20DF9/2020\|EPI_ISL_647589\|20 | ···················· | ········T··········· | ···················· | ···················· | ···················· | ···················· |
| R | 3766 hCoV-19/England/CAMC-B20D26/2020\|EPI_ISL_647590\|20 | ···················· | ···················· | ···················· | ···················· | ···················· | ···················· |
| R | 3767 hCoV-19/England/ALDP-B1A493/2020\|EPI_ISL_647591\|20 | ···················· | ···················· | ···················· | ···················· | ···················· | ···················· |
| R | 3768 hCoV-19/England/CAMC-B2098C/2020\|EPI_ISL_647593\|20 | ···················· | ···················· | ···················· | ···················· | ···················· | ···················· |
| R | 3769 hCoV-19/England/ALDP-B1AA55/2020\|EPI_ISL_647594\|20 | ···················· | ···················· | ···················· | ···················· | ···················· | ···················· |
| R | 3770 hCoV-19/England/CAMC-B20F99/2020\|EPI_ISL_647595\|20 | ···················· | ···················· | ···················· | ···················· | ···················· | ···················· |
| R | 3771 hCoV-19/England/CAMC-B208F8/2020\|EPI_ISL_647596\|20 | ···················· | ···················· | ···················· | ···················· | ···················· | ···················· |
| R | 3772 hCoV-19/England/CAMC-B20B77/2020\|EPI_ISL_647598\|20 | ···················· | ········T··········· | ···················· | ···················· | ···················· | ···················· |
| R | 3773 hCoV-19/England/CAMC-B20227/2020\|EPI_ISL_647599\|20 | ···················· | ···················· | ···················· | ···················· | ···················· | ···················· |
| R | 3774 hCoV-19/England/CAMC-B1FE81/2020\|EPI_ISL_647600\|20 | ···················· | ···················· | ···················· | ···················· | ···················· | ···················· |

| | | | | | | | |
|---|---|---|---|---|---|---|---|
| R | 3775 hCoV-19/England/CAMC-B20737/2020\|EPI_ISL_647601\|20 | ···················· | ···················· | ···················· | ···················· | ···················· | ···················· |
| R | 3776 hCoV-19/England/ALDP-B1A5EB/2020\|EPI_ISL_647603\|20 | ···················· | ···················· | ···················· | ···················· | ···················· | ···················· |
| R | 3777 hCoV-19/England/CAMC-B206D0/2020\|EPI_ISL_647604\|20 | ···················· | ···················· | ···················· | ···················· | ···················· | ···················· |
| R | 3778 hCoV-19/England/CAMC-B20579/2020\|EPI_ISL_647610\|20 | ···················· | ···················· | ···················· | ···················· | ···T················ | ···················· |
| R | 3779 hCoV-19/England/CAMC-B20E05/2020\|EPI_ISL_647611\|20 | ···················· | ···················· | ···················· | ···················· | ···················· | ···················· |
| R | 3780 hCoV-19/England/CAMC-B1FD57/2020\|EPI_ISL_647613\|20 | ···················· | ···················· | ···················· | ···················· | ···················· | ···················· |
| R | 3781 hCoV-19/England/CAMC-B20245/2020\|EPI_ISL_647616\|20 | ···················· | ···················· | ···················· | ···················· | ···················· | ···················· |
| R | 3782 hCoV-19/England/CAMC-B20CA1/2020\|EPI_ISL_647617\|20 | ···················· | ···················· | ···················· | ···················· | ···················· | ···················· |
| R | 3783 hCoV-19/England/ALDP-B1A572/2020\|EPI_ISL_647618\|20 | ···················· | ···················· | ···················· | ···················· | ···················· | ···················· |
| R | 3784 hCoV-19/England/CAMC-B1FDEE/2020\|EPI_ISL_647619\|20 | ···················· | ···················· | ···················· | ···················· | ···················· | ···················· |
| R | 3785 hCoV-19/England/CAMC-B1FE45/2020\|EPI_ISL_647620\|20 | ···················· | ···················· | ···················· | ···················· | ···················· | ···················· |
| R | 3786 hCoV-19/England/CAMC-B21226/2020\|EPI_ISL_647622\|20 | ···················· | ···················· | ···················· | ···················· | ···················· | ···················· |
| R | 3787 hCoV-19/England/CAMC-B20218/2020\|EPI_ISL_647623\|20 | ···················· | ···················· | ···················· | ···················· | ···················· | ···················· |
| R | 3788 hCoV-19/England/ALDP-B1A91C/2020\|EPI_ISL_647624\|20 | ···················· | ···················· | ···················· | ···················· | ···················· | ···················· |
| R | 3789 hCoV-19/England/CAMC-B20290/2020\|EPI_ISL_647625\|20 | ···················· | ···················· | ···················· | ···················· | ···················· | ···················· |
| R | 3790 hCoV-19/England/CAMC-B1FFCA/2020\|EPI_ISL_647626\|20 | ···················· | ···················· | ···················· | ···················· | ···················· | ···················· |
| R | 3791 hCoV-19/USA/CA-ALSR-4195/2020\|EPI_ISL_648880\|2020- | ···················· | ···················· | ···················· | ···················· | ···················· | ···················· |
| R | 3792 hCoV-19/USA/CA-ALSR-4196/2020\|EPI_ISL_648881\|2020- | ···················· | ···················· | ···················· | ···················· | ···················· | ···················· |
| R | 3793 hCoV-19/USA/CA-ALSR-4197/2020\|EPI_ISL_648882\|2020- | ···················· | ·A·················· | ···················· | ···················· | ···················· | ···················· |
| R | 3794 hCoV-19/USA/CA-ALSR-4203/2020\|EPI_ISL_648884\|2020- | ···················· | ·A·················· | ···················· | ···················· | ···················· | ···················· |
| R | 3795 hCoV-19/USA/CA-ALSR-4205/2020\|EPI_ISL_648885\|2020- | ···················· | ·A·················· | ···················· | ···················· | ···················· | ···················· |
| R | 3796 hCoV-19/USA/CA-ALSR-4206/2020\|EPI_ISL_648886\|2020- | ···················· | ·A·················· | ···················· | ···················· | ···················· | ···················· |
| R | 3797 hCoV-19/USA/CA-ALSR-4208/2020\|EPI_ISL_648887\|2020- | ···················· | ·A·················· | ···················· | ···················· | ···················· | ···················· |
| R | 3798 hCoV-19/USA/CA-ALSR-4209/2020\|EPI_ISL_648888\|2020- | ···················· | ·A·················· | ···················· | ···················· | ···················· | ···················· |
| R | 3799 hCoV-19/USA/CA-ALSR-4211/2020\|EPI_ISL_648889\|2020- | ···················· | ···················· | ···················· | ···················· | ···················· | ···················· |
| R | 3800 hCoV-19/USA/CA-ALSR-4213/2020\|EPI_ISL_648890\|2020- | ···················· | ·A·················· | ···················· | ···················· | ···················· | ···················· |
| R | 3801 hCoV-19/USA/CA-ALSR-4216/2020\|EPI_ISL_648891\|2020- | ···················· | ·A·················· | ···················· | ···················· | ···················· | ···················· |
| R | 3802 hCoV-19/USA/CA-ALSR-4217/2020\|EPI_ISL_648892\|2020- | ···················· | ·A·················· | ···················· | ···················· | ···················· | ···················· |
| R | 3803 hCoV-19/USA/CA-ALSR-4218/2020\|EPI_ISL_648893\|2020- | ···················· | ·A·················· | ···················· | ···················· | ···················· | ···················· |
| R | 3804 hCoV-19/USA/CA-ALSR-4220/2020\|EPI_ISL_648894\|2020- | ···················· | ·A·················· | ···················· | ···················· | ···················· | ···················· |
| R | 3805 hCoV-19/USA/CA-ALSR-4223/2020\|EPI_ISL_648896\|2020- | ···················· | ·A·················· | ···················· | ···················· | ···················· | ···················· |
| R | 3806 hCoV-19/USA/CA-ALSR-4224/2020\|EPI_ISL_648897\|2020- | ···················· | ·A·················· | ···················· | ···················· | ···················· | ···················· |
| R | 3807 hCoV-19/USA/CA-ALSR-4229/2020\|EPI_ISL_648899\|2020- | ···················· | ···················· | ···················· | ···················· | ···················· | ···················· |
| R | 3808 hCoV-19/USA/CA-ALSR-4252/2020\|EPI_ISL_648914\|2020- | ···················· | ···················· | ···················· | ···················· | ···················· | ···················· |
| R | 3809 hCoV-19/USA/CA-ALSR-4253/2020\|EPI_ISL_648915\|2020- | ···················· | ···················· | ···················· | ···················· | ···················· | ···················· |
| R | 3810 hCoV-19/USA/CA-ALSR-4254/2020\|EPI_ISL_648916\|2020- | ···················· | ·A·················· | ···················· | ···················· | ···················· | ···················· |
| R | 3811 hCoV-19/USA/CA-ALSR-4256/2020\|EPI_ISL_648918\|2020- | ···················· | ·A·················· | ···················· | ···················· | ···················· | ···················· |
| R | 3812 hCoV-19/USA/CA-ALSR-4259/2020\|EPI_ISL_648919\|2020- | ···················· | ·A·················· | ···················· | ···················· | ···················· | ···················· |
| R | 3813 hCoV-19/USA/CA-ALSR-4260/2020\|EPI_ISL_648920\|2020- | ···················· | ·A·················· | ···················· | ···················· | ···················· | ···················· |
| R | 3814 hCoV-19/USA/CA-ALSR-4261/2020\|EPI_ISL_648921\|2020- | ···················· | ···················· | ···················· | ···················· | ···················· | ···················· |
| R | 3815 hCoV-19/USA/CA-ALSR-4262/2020\|EPI_ISL_648922\|2020- | ···················· | ·A·················· | ···················· | ···················· | ···················· | ···················· |
| R | 3816 hCoV-19/USA/CA-ALSR-4263/2020\|EPI_ISL_648923\|2020- | ···················· | ·············A······ | ···················· | ···················· | ···················· | ···················· |
| R | 3817 hCoV-19/USA/CA-ALSR-4265/2020\|EPI_ISL_648924\|2020- | ···················· | ···················· | ···················· | ···················· | ···················· | ···················· |
| R | 3818 hCoV-19/USA/CA-ALSR-4267/2020\|EPI_ISL_648926\|2020- | ···················· | ·A·················· | ···················· | ···················· | ···················· | ···················· |
| R | 3819 hCoV-19/USA/CA-ALSR-4269/2020\|EPI_ISL_648927\|2020- | ···················· | ·A·················· | ···················· | ···················· | ···················· | ···················· |
| R | 3820 hCoV-19/USA/CA-ALSR-4273/2020\|EPI_ISL_648929\|2020- | ···················· | ·A·················· | ···················· | ···················· | ···················· | ···················· |
| R | 3821 hCoV-19/USA/CA-ALSR-4274/2020\|EPI_ISL_648930\|2020- | ···················· | ·A·················· | ···················· | ···················· | ···················· | ···················· |
| R | 3822 hCoV-19/USA/CA-ALSR-4276/2020\|EPI_ISL_648931\|2020- | ···················· | ·A·················· | ···················· | ···················· | ···················· | ···················· |
| R | 3823 hCoV-19/USA/CA-ALSR-4279/2020\|EPI_ISL_648932\|2020- | ···················· | ···················· | ···················· | ···················· | ···················· | ···················· |
| R | 3824 hCoV-19/USA/CA-ALSR-4280/2020\|EPI_ISL_648933\|2020- | ···················· | ···················· | ···················· | ···················· | ···················· | ···················· |
| R | 3825 hCoV-19/USA/CA-ALSR-4283/2020\|EPI_ISL_648935\|2020- | ···················· | ···················· | ···················· | ···················· | ···················· | ···················· |
| R | 3826 hCoV-19/USA/CA-ALSR-4288/2020\|EPI_ISL_648940\|2020- | ···················· | ···················· | ···················· | ···················· | ···················· | ···················· |
| R | 3827 hCoV-19/USA/CA-ALSR-4289/2020\|EPI_ISL_648941\|2020- | ···················· | ···················· | ···················· | ···················· | ···················· | ···················· |
| R | 3828 hCoV-19/USA/CA-ALSR-4290/2020\|EPI_ISL_648942\|2020- | ···················· | ·A·················· | ···················· | ···················· | ···················· | ···················· |
| R | 3829 hCoV-19/USA/CA-ALSR-4291/2020\|EPI_ISL_648943\|2020- | ···················· | ·A·················· | ···················· | ···················· | ···················· | ···················· |
| R | 3830 hCoV-19/USA/CA-ALSR-4293/2020\|EPI_ISL_648945\|2020- | ···················· | ·A·················· | ···················· | ···················· | ···················· | ···················· |
| R | 3831 hCoV-19/USA/CA-ALSR-4295/2020\|EPI_ISL_648947\|2020- | ········T··········· | ·A·················· | ······G············· | ···················· | ···················· | ···················· |
| R | 3832 hCoV-19/USA/CA-ALSR-4296/2020\|EPI_ISL_648948\|2020- | ···················· | ·A·················· | ···················· | ···················· | ···················· | ···················· |
| R | 3833 hCoV-19/USA/CA-ALSR-4297/2020\|EPI_ISL_648949\|2020- | ···················· | ···················· | ···················· | ···················· | ···················· | ···················· |
| R | 3834 hCoV-19/USA/CA-ALSR-4300/2020\|EPI_ISL_648951\|2020- | ···················· | ···················· | ···················· | ···················· | ···················· | ···················· |
| R | 3835 hCoV-19/USA/CA-ALSR-4302/2020\|EPI_ISL_648952\|2020- | ···················· | ·A·················· | ···················· | ···················· | ···················· | ···················· |
| R | 3836 hCoV-19/USA/CA-ALSR-4303/2020\|EPI_ISL_648953\|2020- | ···················· | ·A·················· | ···················· | ···················· | ···················· | ···················· |
| R | 3837 hCoV-19/USA/CA-ALSR-4304/2020\|EPI_ISL_648954\|2020- | ···················· | ·A·················· | ···················· | ···················· | ···················· | ···················· |
| R | 3838 hCoV-19/USA/CA-ALSR-4305/2020\|EPI_ISL_648955\|2020- | ···················· | ···················· | ···················· | ···················· | ···················· | ···················· |
| R | 3839 hCoV-19/USA/CA-ALSR-4306/2020\|EPI_ISL_648956\|2020- | ···················· | ·A·················· | ···················· | ···················· | ···················· | ···················· |
| R | 3840 hCoV-19/USA/CA-ALSR-4307/2020\|EPI_ISL_648957\|2020- | ···················· | ···················· | ···················· | ···················· | ···················· | ···················· |
| R | 3841 hCoV-19/Ireland/LH-NVRL-73IRL70372/2020\|EPI_ISL_63 | ···················· | ···················· | ···················· | ···················· | ···················· | ···················· |
| R | 3842 hCoV-19/Ireland/LH-NVRL-73IRL41490/2020\|EPI_ISL_63 | ···················· | ···················· | ···················· | ···················· | ···················· | ···················· |
| R | 3843 hCoV-19/Ireland/LH-NVRL-73IRL70315/2020\|EPI_ISL_63 | ···················· | ···················· | ···················· | ···················· | ···················· | ···················· |
| R | 3844 hCoV-19/Ireland/LH-NVRL-73IRL45358/2020\|EPI_ISL_63 | ···················· | ···················· | ···················· | ···················· | ···················· | ···················· |
| R | 3845 hCoV-19/Ireland/LH-NVRL-73IRL41641/2020\|EPI_ISL_63 | ···················· | ···················· | ···················· | ···················· | ···················· | ···················· |
| R | 3846 hCoV-19/Ireland/SO-NVRL-73IRL81470/2020\|EPI_ISL_63 | ···················· | ···················· | ···················· | ···················· | ···················· | ···················· |
| R | 3847 hCoV-19/Ireland/LH-NVRL-73IRL70320/2020\|EPI_ISL_63 | ···················· | ···················· | ···················· | ···················· | ···················· | ···················· |
| R | 3848 hCoV-19/Ireland/D-NVRL-20IRL69600/2020\|EPI_ISL_639 | ···················· | ···················· | ···················· | ···················· | ···················· | ···················· |
| R | 3849 hCoV-19/Ireland/MN-NVRL-20IRL82803/2020\|EPI_ISL_63 | ···················· | ···················· | ···················· | ···················· | ···················· | ···················· |
| R | 3850 hCoV-19/Ireland/MN-NVRL-20IRL82802/2020\|EPI_ISL_63 | ···················· | ···················· | ···················· | ···················· | ···················· | ···················· |
| R | 3851 hCoV-19/Ireland/MN-NVRL-20IRL82789/2020\|EPI_ISL_63 | ···················· | ···················· | ···················· | ···················· | ···················· | ···················· |
| R | 3852 hCoV-19/Ireland/CN-NVRL-20IRL82783/2020\|EPI_ISL_63 | ···················· | ···················· | ···················· | ···················· | ···················· | ···················· |
| R | 3853 hCoV-19/France/ARA93756/2020\|EPI_ISL_640000\|2020-1 | ···················· | ···················· | ···················· | ···················· | ···················· | ···················· |
| R | 3854 hCoV-19/France/ARA97293/2020\|EPI_ISL_640001\|2020-1 | ···················· | ···················· | ···················· | ···················· | ···················· | ···················· |

| | | | | | | | |
|---|---|---|---|---|---|---|---|
| R | 3855 hCoV-19/France/OCC-NRC02765/2020\|EPI_ISL_640002\|20 | ···················· | ···················· | ···················· | ···················· | ···················· | ···················· |
| R | 3856 hCoV-19/France/ARA93309/2020\|EPI_ISL_640003\|2020-1 | ···················· | ···················· | ···················· | ···················· | ···················· | ···················· |
| R | 3857 hCoV-19/France/ARA93996/2020\|EPI_ISL_640004\|2020-1 | ···················· | ···················· | ···················· | ···················· | ···················· | ···················· |
| R | 3858 hCoV-19/France/ARA-NRC02786/2020\|EPI_ISL_640005\|20 | ···················· | ···················· | ···················· | ···················· | ···················· | ···················· |
| R | 3859 hCoV-19/France/OCC-NRC02755/2020\|EPI_ISL_640006\|20 | ···················· | ···················· | ···················· | ···················· | ···················· | ···················· |
| R | 3860 hCoV-19/France/ARA97048/2020\|EPI_ISL_640007\|2020-1 | ···················· | ···················· | ···················· | ···················· | ···················· | ···················· |
| R | 3861 hCoV-19/France/ARA97376/2020\|EPI_ISL_640008\|2020-1 | ···················· | ···················· | ···················· | ···················· | ···················· | ···················· |
| R | 3862 hCoV-19/France/ARA93759/2020\|EPI_ISL_640009\|2020-1 | ···················· | ···················· | ···················· | ···················· | ···················· | ···················· |
| R | 3863 hCoV-19/France/ARA97311/2020\|EPI_ISL_640010\|2020-1 | ···················· | ···················· | ···················· | ···················· | ···················· | ···················· |
| R | 3864 hCoV-19/France/ARA97193/2020\|EPI_ISL_640011\|2020-1 | ···················· | ···················· | ···················· | ···················· | ···················· | ···················· |
| R | 3865 hCoV-19/France/ARA92942/2020\|EPI_ISL_640012\|2020-1 | ···················· | ···················· | ···················· | ···················· | ···················· | ···················· |
| R | 3866 hCoV-19/France/ARA94189/2020\|EPI_ISL_640013\|2020-1 | ···················· | ···················· | ···················· | ···················· | ···················· | ···················· |
| R | 3867 hCoV-19/France/ARA93236/2020\|EPI_ISL_640014\|2020-1 | ···················· | ···················· | ···················· | ···················· | ···················· | ···················· |
| R | 3868 hCoV-19/South Africa/NHLS-UCT-GS-0013/2020\|EPI_ISL | ···················· | ·A·················· | ···················· | ···················· | ···················· | ···················· |
| R | 3869 hCoV-19/South Africa/NHLS-UCT-GS-0014/2020\|EPI_ISL | ···················· | ·A·················· | ···················· | ···················· | ···················· | ···················· |
| R | 3870 hCoV-19/South Africa/NHLS-UCT-GS-0015/2020\|EPI_ISL | ···················· | ·A·················· | ···················· | ···················· | ···················· | ···················· |
| R | 3871 hCoV-19/South Africa/NHLS-UCT-GS-0016/2020\|EPI_ISL | ···················· | ···················· | ···················· | ···················· | ···················· | ···················· |
| R | 3872 hCoV-19/South Africa/NHLS-UCT-GS-0018/2020\|EPI_ISL | ···················· | ···················· | ···················· | ···················· | ···················· | ···················· |
| R | 3873 hCoV-19/South Africa/NHLS-UCT-GS-0021/2020\|EPI_ISL | ···················· | ···················· | ···················· | ···················· | ···················· | ···················· |
| R | 3875 hCoV-19/South Africa/NHLS-UCT-GS-0023/2020\|EPI_ISL | ···················· | ···················· | ···················· | ···················· | ···················· | ···················· |
| R | 3876 hCoV-19/South Africa/NHLS-UCT-GS-0024/2020\|EPI_ISL | ···················· | ·A·················· | ···················· | ···················· | ···················· | ···················· |
| R | 3877 hCoV-19/South Africa/NHLS-UCT-GS-0027/2020\|EPI_ISL | ···················· | ···················· | ···················· | ···················· | ···················· | ···················· |
| R | 3878 hCoV-19/South Africa/NHLS-UCT-GS-0028/2020\|EPI_ISL | ···················· | ·A·················· | ···················· | ···················· | ···················· | ···················· |
| R | 3879 hCoV-19/South Africa/NHLS-UCT-GS-0030/2020\|EPI_ISL | ···················· | ···················· | ···················· | ···················· | ···················· | ···················· |
| R | 3880 hCoV-19/South Africa/NHLS-UCT-GS-0031/2020\|EPI_ISL | ···················· | ·A·················· | ···················· | ···················· | ···················· | ···················· |
| R | 3881 hCoV-19/South Africa/NHLS-UCT-GS-0032/2020\|EPI_ISL | ···················· | ·A·················· | ···················· | ···················· | ···················· | ···················· |
| R | 3882 hCoV-19/South Africa/NHLS-UCT-GS-0034/2020\|EPI_ISL | ···················· | ···················· | ···················· | ···················· | ···················· | ···················· |
| R | 3883 hCoV-19/South Africa/NHLS-UCT-GS-0046/2020\|EPI_ISL | ···················· | ···················· | ···················· | ···················· | ···················· | ···················· |
| R | 3884 hCoV-19/South Africa/NHLS-UCT-GS-0074/2020\|EPI_ISL | ···················· | ···················· | ···················· | ···················· | ···················· | ···················· |
| R | 3885 hCoV-19/South Africa/NHLS-UCT-GS-0081/2020\|EPI_ISL | ···················· | ···················· | ···················· | ···················· | ···················· | ···················· |
| R | 3886 hCoV-19/South Africa/NHLS-UCT-GS-0084/2020\|EPI_ISL | ···················· | ···················· | ···················· | ···················· | ···················· | ···················· |
| R | 3887 hCoV-19/South Africa/NHLS-UCT-GS-1947/2020\|EPI_ISL | ···················· | ···················· | ···················· | ···················· | ···················· | ···················· |
| R | 3888 hCoV-19/South Africa/NHLS-UCT-GS-1955/2020\|EPI_ISL | ···················· | ·A·················· | ···················· | ···················· | ···················· | ···················· |
| R | 3889 hCoV-19/South Africa/NHLS-UCT-GS-1984/2020\|EPI_ISL | ···················· | ···················· | ···················· | ···················· | ···················· | ···················· |
| R | 3890 hCoV-19/South Africa/NHLS-UCT-GS-1990/2020\|EPI_ISL | ···················· | ···················· | ···················· | ···················· | ···················· | ···················· |
| R | 3891 hCoV-19/South Africa/NHLS-UCT-GS-0146/2020\|EPI_ISL | ···················· | ···················· | ···················· | ···················· | ···················· | ···················· |
| R | 3892 hCoV-19/South Africa/NHLS-UCT-GS-0286/2020\|EPI_ISL | ···················· | ···················· | ···················· | ···················· | ···················· | ···················· |
| R | 3893 hCoV-19/South Africa/NHLS-UCT-GS-1433/2020\|EPI_ISL | ·T·················· | ···················· | ···················· | ···················· | ···················· | ···················· |
| R | 3894 hCoV-19/South Africa/NHLS-UCT-GS-0736/2020\|EPI_ISL | ···················· | ·A·················· | ···················· | ···················· | ···················· | ···················· |
| R | 3895 hCoV-19/South Africa/NHLS-UCT-GS-0804/2020\|EPI_ISL | ···················· | ·A·················· | ···················· | ···················· | ···················· | ···················· |
| R | 3896 hCoV-19/South Africa/NHLS-UCT-GS-1246/2020\|EPI_ISL | ···················· | ···················· | ···················· | ···················· | ···················· | ···················· |
| R | 3897 hCoV-19/South Africa/NHLS-UCT-GS-1827/2020\|EPI_ISL | ···················· | ···················· | ···················· | ···················· | ···················· | ···················· |
| R | 3898 hCoV-19/South Africa/NHLS-UCT-GS-2081/2020\|EPI_ISL | ···················· | ···················· | ···················· | ···················· | ···················· | ···················· |
| R | 3899 hCoV-19/South Africa/NHLS-UCT-GS-2426/2020\|EPI_ISL | ···················· | ···················· | ···················· | ···················· | ···················· | ···················· |
| R | 3900 hCoV-19/South Africa/NHLS-UCT-GS-3231/2020\|EPI_ISL | ···················· | ···················· | ···················· | ···················· | ···················· | ···················· |
| R | 3901 hCoV-19/South Africa/NHLS-UCT-GS-2092/2020\|EPI_ISL | ···················· | ···················· | ···················· | ···················· | ···················· | ···················· |
| R | 3904 hCoV-19/Australia/VIC12288/2020\|EPI_ISL_640852\|202 | ···················· | ···················· | ···················· | ···················· | ···················· | ···················· |
| R | 3905 hCoV-19/Australia/VIC16400/2020\|EPI_ISL_640853\|202 | ···················· | ···················· | ···················· | ···················· | ···················· | ···················· |
| R | 3906 hCoV-19/Australia/VIC16408/2020\|EPI_ISL_640854\|202 | ···················· | ···················· | ···················· | ···················· | ···················· | ···················· |
| R | 3907 hCoV-19/Australia/VIC12220/2020\|EPI_ISL_640856\|202 | ···················· | ···················· | ···················· | ···················· | ···················· | ···················· |
| R | 3908 hCoV-19/Australia/VIC12654/2020\|EPI_ISL_640859\|202 | ···················· | ···················· | ···················· | ···················· | ···················· | ···················· |
| R | 3909 hCoV-19/Australia/VIC12714/2020\|EPI_ISL_640862\|202 | ···················· | ···················· | ···················· | ···················· | ···················· | ···················· |
| R | 3910 hCoV-19/Australia/VIC12566/2020\|EPI_ISL_640864\|202 | ···················· | ···················· | ···················· | ···················· | ···················· | ···················· |
| R | 3911 hCoV-19/mink/Denmark/mDK-5/2020\|EPI_ISL_641399\|202 | ···················· | ···················· | ···················· | ···················· | ···················· | ···················· |
| R | 3912 hCoV-19/England/QEUH-B12E06/2020\|EPI_ISL_642656\|20 | ···················· | ·A·················· | ···················· | ···················· | ···················· | ···A················ |
| R | 3913 hCoV-19/England/QEUH-B109AD/2020\|EPI_ISL_642658\|20 | ···················· | ···················· | ···················· | ···················· | ···················· | ···················· |
| R | 3914 hCoV-19/England/QEUH-B1437E/2020\|EPI_ISL_642659\|20 | ···················· | ···················· | ···················· | ···················· | ···················· | ···················· |
| R | 3915 hCoV-19/England/QEUH-B11335/2020\|EPI_ISL_642660\|20 | ···················· | ···················· | ···················· | ···················· | ···················· | ···················· |
| R | 3916 hCoV-19/England/QEUH-B1461B/2020\|EPI_ISL_642661\|20 | ···················· | ···················· | ···················· | ···················· | ···················· | ···················· |
| R | 3917 hCoV-19/England/QEUH-B123BC/2020\|EPI_ISL_642665\|20 | ···················· | ···················· | ···················· | ···················· | ···················· | ···················· |
| R | 3918 hCoV-19/England/QEUH-B1454B/2020\|EPI_ISL_642666\|20 | ···················· | ···················· | ···················· | ···················· | ···················· | ···················· |
| R | 3919 hCoV-19/England/QEUH-B12B96/2020\|EPI_ISL_642667\|20 | ···················· | ···················· | ···················· | ···················· | ···················· | ···················· |
| R | 3920 hCoV-19/England/QEUH-B120E2/2020\|EPI_ISL_642668\|20 | ···················· | ···················· | ···················· | ···················· | ···················· | ···················· |
| R | 3921 hCoV-19/England/QEUH-B0F130/2020\|EPI_ISL_642669\|20 | ···················· | ···················· | ···················· | ···················· | ···················· | ···················· |
| R | 3922 hCoV-19/England/QEUH-B124F5/2020\|EPI_ISL_642670\|20 | ···················· | ···················· | ···················· | ···················· | ···················· | ···················· |
| R | 3923 hCoV-19/England/QEUH-B10934/2020\|EPI_ISL_642671\|20 | ···················· | ···················· | ···················· | ···················· | ···················· | ···················· |
| R | 3924 hCoV-19/England/QEUH-B14499/2020\|EPI_ISL_642672\|20 | ···················· | ···················· | ···················· | ···················· | ···················· | ···················· |
| R | 3925 hCoV-19/England/QEUH-B10A13/2020\|EPI_ISL_642674\|20 | ···················· | ···················· | ···················· | ···················· | ···················· | ···················· |
| R | 3926 hCoV-19/England/QEUH-B126B3/2020\|EPI_ISL_642675\|20 | ···················· | ···················· | ···················· | ···················· | ···················· | ···················· |
| R | 3927 hCoV-19/England/QEUH-B11326/2020\|EPI_ISL_642676\|20 | ···················· | ···················· | ···················· | ···················· | ···················· | ···················· |
| R | 3928 hCoV-19/England/QEUH-B113BD/2020\|EPI_ISL_642677\|20 | ···················· | ···················· | ···················· | ···················· | ······T············· | ···················· |
| R | 3929 hCoV-19/England/QEUH-B1008A/2020\|EPI_ISL_642678\|20 | ···················· | ···················· | ···················· | ···················· | ···················· | ···················· |
| R | 3930 hCoV-19/England/QEUH-B0F14F/2020\|EPI_ISL_642679\|20 | ···················· | ···················· | ···················· | ···················· | ···················· | ···················· |
| R | 3931 hCoV-19/England/QEUH-B0FFEB/2020\|EPI_ISL_642680\|20 | ···················· | ···················· | ···················· | ···················· | ···················· | ···················· |
| R | 3932 hCoV-19/England/QEUH-B10B89/2020\|EPI_ISL_642681\|20 | ···················· | ···················· | ···················· | ···················· | ···················· | ···················· |
| R | 3933 hCoV-19/England/QEUH-B10020/2020\|EPI_ISL_642683\|20 | ···················· | ···················· | ···················· | ···················· | ···················· | ···················· |
| R | 3934 hCoV-19/England/QEUH-B12C1B/2020\|EPI_ISL_642684\|20 | ···················· | ···················· | ···················· | ···················· | ···················· | ···················· |
| R | 3935 hCoV-19/England/QEUH-B14587/2020\|EPI_ISL_642685\|20 | ···················· | ···················· | ···················· | ···················· | ···················· | ···················· |
| R | 3936 hCoV-19/England/QEUH-B12747/2020\|EPI_ISL_642686\|20 | ···················· | ···················· | ···················· | ···················· | ···················· | ···················· |
| R | 3937 hCoV-19/England/QEUH-B12AC6/2020\|EPI_ISL_642687\|20 | ···················· | ·A·················· | ···················· | ···················· | ···················· | ···················· |

R 3938 hCoV-19/England/QEUH-B12B87/2020|EPI_ISL_642688|20 ···················· ···················· ····················· ···················· ···················· ····················
R 3939 hCoV-19/England/QEUH-B1262C/2020|EPI_ISL_642689|20 ···················· ···················· ····················· ···················· ···················· ····················
R 3940 hCoV-19/England/QEUH-B12E8E/2020|EPI_ISL_642690|20 ···················· ···················· ····················· ···················· ···················· ····················
R 3941 hCoV-19/England/QEUH-B0F1F4/2020|EPI_ISL_642691|20 ···················· ···················· ····················· ···················· ···················· ····················
R 3942 hCoV-19/England/QEUH-B1257A/2020|EPI_ISL_642692|20 ···················· ···················· ····················· ···················· ···················· ····················
R 3943 hCoV-19/England/QEUH-B112A1/2020|EPI_ISL_642693|20 ···················· ···················· ····················· ···················· ···················· ····················
R 3944 hCoV-19/England/QEUH-B123AD/2020|EPI_ISL_642695|20 ···················· ···················· ····················· ···················· ···················· ····················
R 3945 hCoV-19/England/QEUH-B1139F/2020|EPI_ISL_642696|20 ···················· ···················· ····················· ···················· ······T············· ····················
R 3946 hCoV-19/England/QEUH-B12E33/2020|EPI_ISL_642697|20 ···················· ···················· ····················· ···················· ···················· ····················
R 3947 hCoV-19/England/QEUH-B14341/2020|EPI_ISL_642699|20 ···················· ···················· ····················· ···················· ······T············· ····················
R 3948 hCoV-19/England/QEUH-B1263B/2020|EPI_ISL_642700|20 ···················· ···················· ····················· ···················· ···················· ····················
R 3949 hCoV-19/England/QEUH-B10B5C/2020|EPI_ISL_642701|20 ···················· ···················· ····················· ···················· ···················· ····················
R 3950 hCoV-19/England/QEUH-B12C57/2020|EPI_ISL_642702|20 ···················· ···················· ····················· ···················· ···················· ····················
R 3951 hCoV-19/England/QEUH-B1203D/2020|EPI_ISL_642703|20 ···················· ···················· ····················· ···················· ···················· ····················
R 3952 hCoV-19/England/QEUH-B1202E/2020|EPI_ISL_642705|20 ···················· ···················· ····················· ···················· ···················· ····················
R 3953 hCoV-19/England/QEUH-B0F16D/2020|EPI_ISL_642706|20 ···················· ···················· ····················· ···················· ···················· ····················
R 3954 hCoV-19/England/QEUH-B126FF/2020|EPI_ISL_642707|20 ···················· ·A·················· ····················· ···················· ···················· ···A················
R 3955 hCoV-19/England/QEUH-B109BC/2020|EPI_ISL_642708|20 ···················· ···················· ····················· ···················· ···················· ····················
R 3956 hCoV-19/England/QEUH-B120B5/2020|EPI_ISL_642709|20 ···················· ···················· ····················· ···················· ···················· ····················
R 3957 hCoV-19/England/MILK-ACD25A/2020|EPI_ISL_643986|20 ···················· ···················· ····················· ···················· ···················· ····················
R 3958 hCoV-19/England/MILK-AD0139/2020|EPI_ISL_643987|20 ···················· ···················· ····················· ···················· ···················· ····················
R 3959 hCoV-19/England/MILK-AC7BA0/2020|EPI_ISL_643988|20 ···················· ···················· ····················· ···················· ···················· ····················
R 3960 hCoV-19/England/MILK-AD00B4/2020|EPI_ISL_643989|20 ···················· ···················· ····················· ···················· ···················· ····················
R 3961 hCoV-19/England/MILK-ACD81C/2020|EPI_ISL_643991|20 ···················· ···················· ····················· ···················· ···················· ····················
R 3962 hCoV-19/Scotland/QEUH-AD1C0A/2020|EPI_ISL_643992|2 ···················· ···················· ····················· ···················· ···················· ····················
R 3963 hCoV-19/Scotland/QEUH-AD4A76/2020|EPI_ISL_643994|2 ···················· ···················· ····················· ···················· ···················· ····················
R 3964 hCoV-19/Scotland/QEUH-AD4A94/2020|EPI_ISL_643995|2 ···················· ···················· ····················· ···················· ···················· ····················
R 3965 hCoV-19/Scotland/QEUH-AD1C55/2020|EPI_ISL_643997|2 ···················· ···················· ····················· ···················· ···················· ····················
R 3966 hCoV-19/Scotland/QEUH-AD02FA/2020|EPI_ISL_643998|2 ···················· ···················· ····················· ···················· ···················· ····················
R 3967 hCoV-19/Scotland/QEUH-AD1BFE/2020|EPI_ISL_643999|2 ···················· ···················· ····················· ···················· ···················· ····················
R 3968 hCoV-19/England/QEUH-B2E089/2020|EPI_ISL_646200|20 ···················· ···················· ····················· ···················· ···················· ····················
R 3969 hCoV-19/England/QEUH-B31171/2020|EPI_ISL_646201|20 ···················· ···················· ····················· ···················· ···················· ····················
R 3970 hCoV-19/England/QEUH-B311BD/2020|EPI_ISL_646202|20 ···················· ···················· ····················· ···················· ···················· ····················
R 3971 hCoV-19/England/QEUH-B31F4A/2020|EPI_ISL_646203|20 ···················· ···················· ····················· ···················· ···················· ····················
R 3972 hCoV-19/England/ALDP-B23DF6/2020|EPI_ISL_646204|20 ···················· ···················· ····················· ···················· ···················· ····················
R 3973 hCoV-19/England/QEUH-B32170/2020|EPI_ISL_646205|20 ···················· ···················· ····················· ···················· ···················· ····················
R 3974 hCoV-19/England/QEUH-B320A0/2020|EPI_ISL_646206|20 ···················· ···················· ····················· ···················· ···················· ····················
R 3975 hCoV-19/England/ALDP-B23B29/2020|EPI_ISL_646207|20 ···················· ···················· ····················· ···················· ···················· ····················
R 3976 hCoV-19/England/ALDP-B23D9C/2020|EPI_ISL_646208|20 ···················· ···················· ····················· ···················· ···················· ····················
R 3977 hCoV-19/England/QEUH-B32028/2020|EPI_ISL_646209|20 ···················· ···················· ····················· ···················· ···················· ····················
R 3978 hCoV-19/England/QEUH-B31117/2020|EPI_ISL_646211|20 ···················· ···················· ····················· ···················· ···················· ····················
R 3979 hCoV-19/England/ALDP-B24092/2020|EPI_ISL_646212|20 ············T······· ·A·················· ····················· ···················· ···················· ····················
R 3980 hCoV-19/England/QEUH-B31E10/2020|EPI_ISL_646213|20 ···················· ···················· ····················· ···················· ···················· ····················
R 3981 hCoV-19/England/QEUH-B2F747/2020|EPI_ISL_646214|20 ···················· ···················· ····················· ···················· ···················· ····················
R 3982 hCoV-19/England/ALDP-B24038/2020|EPI_ISL_646215|20 ···················· ···················· ····················· ···················· ···················· ····················
R 3983 hCoV-19/England/QEUH-B31E98/2020|EPI_ISL_646216|20 ···················· ···················· ····················· ···················· ···················· ····················
R 3984 hCoV-19/England/QEUH-B318C7/2020|EPI_ISL_646217|20 ···················· ···················· ····················· ···················· ···················· ····················
R 3985 hCoV-19/England/QEUH-B318F4/2020|EPI_ISL_646218|20 ···················· ···················· ····················· ···················· ···················· ····················
R 3986 hCoV-19/England/CAMC-B20E32/2020|EPI_ISL_647501|20 ···················· ···················· ····················· ···················· ···················· ····················
R 3987 hCoV-19/England/CAMC-B20ED8/2020|EPI_ISL_647502|20 ···················· ···················· ····················· ···················· ···················· ····················
R 3988 hCoV-19/England/CAMC-B211CF/2020|EPI_ISL_647505|20 ···················· ···················· ····················· ···················· ···················· ····················
R 3989 hCoV-19/England/ALDP-B1AA64/2020|EPI_ISL_647506|20 ···················· ···················· ····················· ···················· ···················· ····················
R 3990 hCoV-19/England/CAMC-B20A10/2020|EPI_ISL_647508|20 ···················· ···················· ····················· ···················· ···················· ····················
R 3991 hCoV-19/England/CAMC-B1FEAF/2020|EPI_ISL_647509|20 ···················· ···················· ····················· ···················· ···················· ····················
R 3992 hCoV-19/England/CAMC-B1FF9D/2020|EPI_ISL_647510|20 ···················· ···················· ····················· ···················· ···················· ····················
R 3993 hCoV-19/England/ALDP-B1A7D6/2020|EPI_ISL_647511|20 ···················· ·A·················· ····················· ···················· ···················· ····················
R 3994 hCoV-19/England/ALDP-B1A84C/2020|EPI_ISL_647513|20 ···················· ···················· ····················· ···················· ···················· ····················
R 3995 hCoV-19/England/CAMC-B1FE36/2020|EPI_ISL_647514|20 ···················· ···················· ····················· ···················· ···················· ····················
R 3996 hCoV-19/England/CAMC-B1FE54/2020|EPI_ISL_647515|20 ···················· ···················· ····················· ···················· ···················· ····················
R 3997 hCoV-19/England/CAMC-B2002D/2020|EPI_ISL_647517|20 ···················· ···················· ····················· ···················· ···················· ····················
R 3998 hCoV-19/England/CAMC-B1FC1E/2020|EPI_ISL_647519|20 ···················· ···················· ····················· ···················· ···················· ····················
R 3999 hCoV-19/England/CAMC-B1FD75/2020|EPI_ISL_647520|20 ···················· ···················· ····················· ···················· ···················· ····················
R 4000 hCoV-19/England/CAMC-B20EBA/2020|EPI_ISL_647521|20 ···················· ···················· ····················· ···················· ···················· ····················
R 4001 hCoV-19/England/CAMC-B1FC3C/2020|EPI_ISL_647522|20 ···················· ···················· ····················· ···················· ···················· ····················
R 4002 hCoV-19/England/CAMC-B21138/2020|EPI_ISL_647523|20 ···················· ···················· ····················· ···················· ···················· ····················
R 4003 hCoV-19/England/CAMC-B1FB7B/2020|EPI_ISL_647524|20 ···················· ···················· ····················· ···················· ···················· ····················
R 4004 hCoV-19/England/CAMC-B20B59/2020|EPI_ISL_647525|20 ···················· ···················· ····················· ···················· ······T············· ····················
R 4005 hCoV-19/England/ALDP-B1A5AF/2020|EPI_ISL_647526|20 ···················· ···················· ····················· ···················· ···················· ····················
R 4006 hCoV-19/England/CAMC-B20D80/2020|EPI_ISL_647527|20 ···················· ···················· ····················· ···················· ······T············· ····················
R 4007 hCoV-19/England/ALDP-B1A9EF/2020|EPI_ISL_647528|20 ···················· ···················· ····················· ···················· ···················· ····················
R 4008 hCoV-19/England/CAMC-B20C38/2020|EPI_ISL_647529|20 ···················· ···················· ····················· ···················· ···················· ····················
R 4009 hCoV-19/England/CAMC-B2152D/2020|EPI_ISL_647530|20 ···················· ···················· ····················· ···················· ···················· ····················
R 4010 hCoV-19/England/CAMC-B1FD39/2020|EPI_ISL_647531|20 ···················· ···················· ····················· ···················· ···················· ····················
R 4011 hCoV-19/England/CAMC-B20D44/2020|EPI_ISL_647532|20 ···················· ···················· ····················· ···················· ···················· ····················
R 4012 hCoV-19/England/CAMC-B20E9C/2020|EPI_ISL_647533|20 ···················· ···················· ····················· ···················· ···················· ····················
R 4013 hCoV-19/England/CAMC-B20AB6/2020|EPI_ISL_647534|20 ···················· ···················· ····················· ···················· ···················· ····················
R 4014 hCoV-19/England/ALDP-B1A9C1/2020|EPI_ISL_647535|20 ···················· ···················· ····················· ···················· ···················· ····················
R 4015 hCoV-19/England/ALDP-B1A976/2020|EPI_ISL_647536|20 ···················· ···················· ····················· ···················· ···················· ····················
R 4016 hCoV-19/England/CAMC-B20209/2020|EPI_ISL_647537|20 ···················· ···················· ····················· ···················· ···················· ····················
R 4017 hCoV-19/England/CAMC-B21596/2020|EPI_ISL_647538|20 ···················· ···················· ····················· ···················· ···················· ····················

```
R 4018 hCoV-19/England/CAMC-B20588/2020|EPI_ISL_647539|20 ···················· ···················· ····················· ···················· ····T··············· ····················
R 4019 hCoV-19/England/CAMC-B205A6/2020|EPI_ISL_647540|20 ···················· ·A·················· ····················· ···················· ···················· ···A················
R 4020 hCoV-19/England/ALDP-B1AA46/2020|EPI_ISL_647541|20 ···················· ·A·················· ····················· ···················· ···················· ····················
R 4021 hCoV-19/England/CAMC-B1FC78/2020|EPI_ISL_647542|20 ···················· ·A·······A·········· ····················· ···················· ···················· ····················
R 4022 hCoV-19/England/CAMC-B20BA4/2020|EPI_ISL_647543|20 ···················· ···················· ····················· ···················· ···················· ····················
R 4023 hCoV-19/England/CAMC-B20F02/2020|EPI_ISL_647544|20 ···················· ···················· ····················· ···················· ···················· ····················
R 4024 hCoV-19/England/CAMC-B20236/2020|EPI_ISL_647545|20 ···················· ·A·················· ····················· ···················· ···················· ···A················
R 4025 hCoV-19/England/ALDP-B1A545/2020|EPI_ISL_647546|20 ···················· ···················· ····················· ···················· ···················· ····················
R 4026 hCoV-19/England/CAMC-B20A7A/2020|EPI_ISL_647547|20 ···················· ···················· ····················· ···················· ···················· ····················
R 4027 hCoV-19/England/CAMC-B2153C/2020|EPI_ISL_647548|20 ···················· ···················· ····················· ···················· ···················· ····················
R 4028 hCoV-19/Singapore/1313/2020|EPI_ISL_648800|2020-05 ···················· ···················· ····················· ···················· ···················· ····················
R 4029 hCoV-19/Singapore/1314/2020|EPI_ISL_648801|2020-05 ···················· ···················· ····················· ···················· ···················· ····················
R 4030 hCoV-19/Singapore/1315/2020|EPI_ISL_648802|2020-05 ···················· ···················· ····················· ···················· ···················· ····················
R 4031 hCoV-19/Singapore/1316/2020|EPI_ISL_648803|2020-05 ···················· ···················· ····················· ···················· ···················· ····················
R 4032 hCoV-19/Singapore/320nan/2020|EPI_ISL_648804|2020- ···················· ···················· ····················· ···················· ···················· ····················
R 4033 hCoV-19/Singapore/1317/2020|EPI_ISL_648805|2020-05 ···················· ···················· ····················· ···················· ···················· ····················
R 4034 hCoV-19/Singapore/1318/2020|EPI_ISL_648806|2020-05 ···················· ···················· ····················· ···················· ···················· ····················
R 4035 hCoV-19/Singapore/1319/2020|EPI_ISL_648807|2020-05 ···················· ···················· ····················· ···················· ···················· ····················
R 4036 hCoV-19/Singapore/1320/2020|EPI_ISL_648808|2020-05 ···················· ···················· ····················· ···················· ···················· ····················
R 4037 hCoV-19/Singapore/1321/2020|EPI_ISL_648809|2020-05 ···················· ···················· ····················· ···················· ···················· ····················
R 4038 hCoV-19/Singapore/1322/2020|EPI_ISL_648810|2020-05 ···················· ···················· ····················· ···················· ···················· ····················
R 4039 hCoV-19/Singapore/1323/2020|EPI_ISL_648811|2020-05 ···················· ···················· ····················· ···················· ···················· ····················
R 4040 hCoV-19/Singapore/1324/2020|EPI_ISL_648812|2020-05 ···················· ···················· ····················· ···················· ···················· ····················
R 4041 hCoV-19/Singapore/1325/2020|EPI_ISL_648813|2020-05 ···················· ···················· ····················· ···················· ···················· ····················
R 4042 hCoV-19/Singapore/1326/2020|EPI_ISL_648814|2020-05 ···················· ···················· ····················· ···················· ···················· ····················
R 4043 hCoV-19/Singapore/547nan/2020|EPI_ISL_648815|2020- ···················· ···················· ····················· ···················· ···················· ····················
R 4044 hCoV-19/Singapore/1327/2020|EPI_ISL_648816|2020-05 ···················· ···················· ····················· ···················· ···················· ····················
R 4045 hCoV-19/Singapore/1328/2020|EPI_ISL_648817|2020-05 ···················· ···················· ····················· ···················· ···················· ····················
R 4046 hCoV-19/Singapore/1329/2020|EPI_ISL_648818|2020-06 ···················· ···················· ····················· ···················· ···················· ····················
R 4047 hCoV-19/Singapore/1330/2020|EPI_ISL_648819|2020-06 ···················· ···················· ····················· ···················· ···················· ····················
R 4048 hCoV-19/Singapore/1331/2020|EPI_ISL_648820|2020-06 ···················· ···················· ····················· ···················· ···················· ····················
R 4049 hCoV-19/USA/MT-RML-22/2020|EPI_ISL_648822|2020-08- ···················· ··C················· ····················· ···················· ···················· ····················
R 4050 hCoV-19/USA/CA-ALSR-4099/2020|EPI_ISL_648823|2020- ···················· ·A·················· ····················· ···················· ···················· ····················
R 4051 hCoV-19/USA/CA-ALSR-4102/2020|EPI_ISL_648824|2020- ···················· ·A·················· ····················· ···················· ···················· ····················
R 4052 hCoV-19/USA/CA-ALSR-4103/2020|EPI_ISL_648825|2020- ···················· ·A·················· ····················· ···················· ···················· ····················
R 4053 hCoV-19/USA/CA-ALSR-4106/2020|EPI_ISL_648826|2020- ···················· ·A·················· ····················· ···················· ···················· ····················
R 4054 hCoV-19/USA/CA-ALSR-4107/2020|EPI_ISL_648827|2020- ···················· ·A·················· ····················· ···················· ···················· ····················
R 4055 hCoV-19/USA/CA-ALSR-4108/2020|EPI_ISL_648828|2020- ···················· ·A·················· ····················· ···················· ···················· ····················
R 4056 hCoV-19/USA/CA-ALSR-4116/2020|EPI_ISL_648830|2020- ···················· ·A·················· ····················· ···················· ···················· ····················
R 4057 hCoV-19/USA/CA-ALSR-4123/2020|EPI_ISL_648832|2020- ···················· ·A·················· ····················· ···················· ···················· ····················
R 4058 hCoV-19/USA/CA-ALSR-4124/2020|EPI_ISL_648833|2020- ···················· ·A·················· ····················· ···················· ···················· ····················
R 4059 hCoV-19/USA/CA-ALSR-4125/2020|EPI_ISL_648834|2020- ···················· ·A·················· ····················· ···················· ···················· ····················
R 4060 hCoV-19/USA/CA-ALSR-4126/2020|EPI_ISL_648835|2020- ···················· ···················· ····················· ···················· ···················· ····················
R 4061 hCoV-19/USA/CA-ALSR-4128/2020|EPI_ISL_648836|2020- ···················· ·A·················· ····················· ···················· ···················· ····················
R 4062 hCoV-19/USA/CA-ALSR-4129/2020|EPI_ISL_648837|2020- ···················· ···················· ····················· ···················· ···················· ····················
R 4063 hCoV-19/USA/CA-ALSR-4130/2020|EPI_ISL_648838|2020- ···················· ·A·················· ····················· ···················· ···················· ····················
R 4064 hCoV-19/USA/CA-ALSR-4131/2020|EPI_ISL_648839|2020- ···················· ·A·················· ····················· ···················· ···················· ····················
R 4065 hCoV-19/USA/CA-ALSR-4133/2020|EPI_ISL_648840|2020- ···················· ·A·················· ····················· ···················· ···················· ····················
R 4066 hCoV-19/USA/CA-ALSR-4134/2020|EPI_ISL_648841|2020- ···················· ·A·················· ····················· ···················· ···················· ····················
R 4067 hCoV-19/USA/CA-ALSR-4139/2020|EPI_ISL_648843|2020- ···················· ·A·················· ····················· ···················· ···················· ····················
R 4068 hCoV-19/USA/CA-ALSR-4140/2020|EPI_ISL_648844|2020- ···················· ·A·················· ····················· ···················· ···················· ····················
R 4069 hCoV-19/USA/CA-ALSR-4141/2020|EPI_ISL_648845|2020- ···················· ·A·················· ····················· ···················· ···················· ····················
R 4070 hCoV-19/USA/CA-ALSR-4142/2020|EPI_ISL_648846|2020- ···················· ·A·················· ····················· ···················· ···················· ····················
R 4071 hCoV-19/USA/CA-ALSR-4143/2020|EPI_ISL_648847|2020- ···················· ·A·················· ····················· ···················· ···················· ····················
R 4072 hCoV-19/USA/CA-ALSR-4144/2020|EPI_ISL_648848|2020- ···················· ·A·················· ····················· ···················· ···················· ····················
R 4073 hCoV-19/USA/CA-ALSR-4146/2020|EPI_ISL_648850|2020- ···················· ·A·················· ····················· ···················· ···················· ····················
R 4074 hCoV-19/USA/CA-ALSR-4148/2020|EPI_ISL_648851|2020- ···················· ·A·················· ····················· ···················· ···················· ····················
R 4075 hCoV-19/USA/CA-ALSR-4149/2020|EPI_ISL_648852|2020- ···················· ···················· ····················· ···················· ···················· ····················
R 4076 hCoV-19/USA/CA-ALSR-4150/2020|EPI_ISL_648853|2020- ···················· ·A·················· ····················· ···················· ···················· ····················
R 4077 hCoV-19/USA/CA-ALSR-4151/2020|EPI_ISL_648854|2020- ···················· ·A·················· ····················· ···················· ···················· ····················
R 4078 hCoV-19/USA/CA-ALSR-4152/2020|EPI_ISL_648855|2020- ···················· ·A·················· ····················· ···················· ···················· ····················
R 4079 hCoV-19/USA/CA-ALSR-4155/2020|EPI_ISL_648856|2020- ···················· ·A·················· ····················· ···················· ···················· ····················
R 4080 hCoV-19/USA/CA-ALSR-4156/2020|EPI_ISL_648857|2020- ···················· ·A·················· ····················· ···················· ···················· ····················
R 4081 hCoV-19/USA/CA-ALSR-4157/2020|EPI_ISL_648858|2020- ···················· ·A·················· ····················· ···················· ···················· ····················
R 4082 hCoV-19/USA/CA-ALSR-4164/2020|EPI_ISL_648859|2020- ···················· ·A·················· ····················· ···················· ···················· ····················
R 4083 hCoV-19/USA/CA-ALSR-4171/2020|EPI_ISL_648862|2020- ···················· ·A·················· ····················· ···················· ···················· ····················
R 4084 hCoV-19/USA/CA-ALSR-4172/2020|EPI_ISL_648863|2020- ···················· ·A·················· ····················· ···················· ···················· ····················
R 4085 hCoV-19/USA/CA-ALSR-4173/2020|EPI_ISL_648864|2020- ···················· ·A·················· ····················· ···················· ···················· ····················
R 4086 hCoV-19/USA/CA-ALSR-4176/2020|EPI_ISL_648865|2020- ···················· ·A·················· ····················· ···················· ···················· ····················
R 4087 hCoV-19/USA/CA-ALSR-4177/2020|EPI_ISL_648866|2020- ···················· ·A·················· ····················· ···················· ···················· ····················
R 4088 hCoV-19/USA/CA-ALSR-4182/2020|EPI_ISL_648868|2020- ···················· ···················· ···········T········· ···················· ···················· ····················
R 4089 hCoV-19/USA/CA-ALSR-4184/2020|EPI_ISL_648870|2020- ···················· ·A·················· ····················· ···················· ···················· ····················
R 4090 hCoV-19/USA/CA-ALSR-4185/2020|EPI_ISL_648871|2020- ···················· ·A·················· ····················· ···················· ···················· ····················
R 4091 hCoV-19/USA/CA-ALSR-4186/2020|EPI_ISL_648872|2020- ···················· ·A·················· ····················· ···················· ···················· ····················
R 4092 hCoV-19/USA/CA-ALSR-4188/2020|EPI_ISL_648874|2020- ···················· ·A·················· ····················· ···················· ···················· ····················
R 4093 hCoV-19/USA/CA-ALSR-4189/2020|EPI_ISL_648875|2020- ···················· ·A·················· ····················· ···················· ···················· ····················
R 4094 hCoV-19/USA/CA-ALSR-4190/2020|EPI_ISL_648876|2020- ···················· ·A·················· ····················· ···················· ···················· ····················
R 4095 hCoV-19/USA/CA-ALSR-4191/2020|EPI_ISL_648877|2020- ···················· ·A·················· ····················· ···················· ···················· ····················
R 4096 hCoV-19/USA/CA-ALSR-4192/2020|EPI_ISL_648878|2020- ···················· ·A·················· ····················· ···················· ···················· ····················
R 4097 hCoV-19/Australia/QLD1215/2020|EPI_ISL_639806|2020 ···················· ···················· ····················· ···················· ···················· ····················
```

| | | | | | | | |
|---|---|---|---|---|---|---|---|
| R | 4098 hCoV-19/Australia/QLD1216/2020\|EPI_ISL_639807\|2020 | ···················· | ···················· | ····················· | ···················· | ···················· | ···················· |
| R | 4099 hCoV-19/Australia/QLD1217/2020\|EPI_ISL_639808\|2020 | ···················· | ···················· | ····················· | ···················· | ···················· | ···················· |
| R | 4100 hCoV-19/Australia/QLD1218/2020\|EPI_ISL_639809\|2020 | ···················· | ···················· | ····················· | ···················· | ···················· | ···················· |
| R | 4101 hCoV-19/Australia/QLD1219/2020\|EPI_ISL_639810\|2020 | ···················· | ···················· | ····················· | ···················· | ···················· | ···················· |
| R | 4102 hCoV-19/Australia/QLD1220/2020\|EPI_ISL_639811\|2020 | ···················· | ···················· | ····················· | ···················· | ···················· | ···················· |
| R | 4103 hCoV-19/Australia/QLD1221/2020\|EPI_ISL_639812\|2020 | ···················· | ···················· | ····················· | ···················· | ···················· | ···················· |
| R | 4104 hCoV-19/Australia/QLD1222/2020\|EPI_ISL_639813\|2020 | ···················· | ···················· | ····················· | ···················· | ···················· | ···················· |
| R | 4105 hCoV-19/Australia/QLD1223/2020\|EPI_ISL_639814\|2020 | ···················· | ···················· | ····················· | ···················· | ···················· | ···················· |
| R | 4106 hCoV-19/Australia/QLD1224/2020\|EPI_ISL_639815\|2020 | ···················· | ···················· | ····················· | ···················· | ···················· | ···················· |
| R | 4107 hCoV-19/Australia/QLD1227/2020\|EPI_ISL_639816\|2020 | ···················· | ···················· | ····················· | ···················· | ···················· | ···················· |
| R | 4108 hCoV-19/Australia/QLD1228/2020\|EPI_ISL_639817\|2020 | ···················· | ···················· | ····················· | ···················· | ···················· | ···················· |
| R | 4109 hCoV-19/Australia/QLD1229/2020\|EPI_ISL_639818\|2020 | ···················· | ···················· | ····················· | ···················· | ···················· | ···················· |
| R | 4110 hCoV-19/Australia/QLD1231/2020\|EPI_ISL_639819\|2020 | ···················· | ···········A········ | ····················· | ···················· | ···················· | ···················· |
| R | 4111 hCoV-19/Australia/QLD1236/2020\|EPI_ISL_639820\|2020 | ···················· | ···················· | ····················· | ···················· | ···················· | ···················· |
| R | 4112 hCoV-19/England/204393874/2020\|EPI_ISL_639821\|2020 | ···················· | ···················· | ····················· | ···················· | ···················· | ···················· |
| R | 4113 hCoV-19/England/204490653/2020\|EPI_ISL_639822\|2020 | ···················· | ···················· | ····················· | ···················· | ···················· | ···················· |
| R | 4114 hCoV-19/Ireland/LH-NVRL-73IRL70373/2020\|EPI_ISL_63 | ···················· | ···················· | ····················· | ···················· | ···················· | ···················· |
| R | 4115 hCoV-19/Ireland/SO-NVRL-73IRL06905/2020\|EPI_ISL_63 | ···················· | ···················· | ····················· | ···················· | ···················· | ···················· |
| R | 4116 hCoV-19/Ireland/LH-NVRL-73IRL40279/2020\|EPI_ISL_63 | ···················· | ···················· | ····················· | ···················· | ···················· | ···················· |
| R | 4117 hCoV-19/Ireland/LH-NVRL-73IRL70403/2020\|EPI_ISL_63 | ···················· | ···················· | ····················· | ···················· | ···················· | ···················· |
| R | 4118 hCoV-19/Ireland/LH-NVRL-73IRL70416/2020\|EPI_ISL_63 | ···················· | ·A·················· | ····················· | ···················· | ···················· | ···A················ |
| R | 4119 hCoV-19/Ireland/MN-NVRL-73IRL72912/2020\|EPI_ISL_63 | ···················· | ···················· | ····················· | ···················· | ···················· | ···················· |
| R | 4120 hCoV-19/Ireland/SO-NVRL-73IRL20048/2020\|EPI_ISL_63 | ···················· | ···················· | ····················· | ···················· | ···················· | ···················· |
| R | 4121 hCoV-19/Ireland/SO-NVRL-73IRL72886/2020\|EPI_ISL_63 | ···················· | ···················· | ····················· | ···················· | ···················· | ···················· |
| R | 4122 hCoV-19/Ireland/MN-NVRL-73IRL72925/2020\|EPI_ISL_63 | ···················· | ···················· | ····················· | ···················· | ···················· | ···················· |
| R | 4123 hCoV-19/Ireland/SO-NVRL-73IRL20020/2020\|EPI_ISL_63 | ···················· | ···················· | ····················· | ···················· | ···················· | ···················· |
| R | 4124 hCoV-19/Ireland/SO-NVRL-73IRL20015/2020\|EPI_ISL_63 | ···················· | ···················· | ····················· | ···················· | ···················· | ···················· |
| R | 4125 hCoV-19/Ireland/LH-NVRL-73IRL41576/2020\|EPI_ISL_63 | ···················· | ···················· | ····················· | ···················· | ···················· | ···················· |
| R | 4126 hCoV-19/Ireland/MH-NVRL-76IRL26088/2020\|EPI_ISL_63 | ···················· | ···················· | ····················· | ···················· | ···················· | ···················· |
| R | 4127 hCoV-19/Ireland/LH-NVRL-73IRL70375/2020\|EPI_ISL_63 | ···················· | ···················· | ····················· | ···················· | ···················· | ···················· |
| R | 4128 hCoV-19/Ireland/MH-NVRL-76IRL26957/2020\|EPI_ISL_63 | ···················· | ···················· | ····················· | ···················· | ···················· | ···················· |
| R | 4129 hCoV-19/Ireland/SO-NVRL-73IRL69623/2020\|EPI_ISL_63 | ···················· | ···················· | ····················· | ···················· | ···················· | ···················· |
| R | 4130 hCoV-19/Ireland/SO-NVRL-73IRL06915/2020\|EPI_ISL_63 | ···················· | ···················· | ····················· | ···················· | ···················· | ···················· |
| R | 4131 hCoV-19/Ireland/SO-NVRL-73IRL69646/2020\|EPI_ISL_63 | ···················· | ···················· | ····················· | ···················· | ···················· | ···················· |
| R | 4132 hCoV-19/Ireland/SO-NVRL-73IRL69650/2020\|EPI_ISL_63 | ···················· | ···················· | ····················· | ···················· | ···················· | ···················· |
| R | 4133 hCoV-19/Ireland/LH-NVRL-73IRL70326/2020\|EPI_ISL_63 | ···················· | ·A·················· | ····················· | ···················· | ···················· | ···A················ |
| R | 4134 hCoV-19/Ireland/LH-NVRL-73IRL41660/2020\|EPI_ISL_63 | ···················· | ···················· | ····················· | ···················· | ···················· | ···················· |
| R | 4135 hCoV-19/Ireland/LH-NVRL-73IRL41493/2020\|EPI_ISL_63 | ···················· | ···················· | ····················· | ···················· | ···················· | ···················· |
| R | 4136 hCoV-19/Ireland/SO-NVRL-73IRL81508/2020\|EPI_ISL_63 | ···················· | ···················· | ····················· | ···················· | ···················· | ···················· |
| R | 4137 hCoV-19/Ireland/LH-NVRL-73IRL70361/2020\|EPI_ISL_63 | ···················· | ···················· | ····················· | ···················· | ···················· | ···················· |
| R | 4138 hCoV-19/Ireland/LH-NVRL-73IRL40264/2020\|EPI_ISL_63 | ···················· | ···················· | ····················· | ···················· | ···················· | ···················· |
| R | 4139 hCoV-19/Ireland/LH-NVRL-73IRL40251/2020\|EPI_ISL_63 | ···················· | ···················· | ····················· | ···················· | ···················· | ···················· |
| R | 4140 hCoV-19/Ireland/CN-NVRL-20IRL82775/2020\|EPI_ISL_63 | ···················· | ···················· | ····················· | ···················· | ···················· | ···················· |
| R | 4141 hCoV-19/Ireland/CN-NVRL-20IRL82733/2020\|EPI_ISL_63 | ···················· | ···················· | ····················· | ···················· | ···················· | ···················· |
| R | 4142 hCoV-19/Ireland/CN-NVRL-20IRL82714/2020\|EPI_ISL_63 | ···················· | ···················· | ····················· | ···················· | ···················· | ···················· |
| R | 4143 hCoV-19/Ireland/CN-NVRL-20IRL82702/2020\|EPI_ISL_63 | ···················· | ···················· | ····················· | ···················· | ···················· | ···················· |
| R | 4144 hCoV-19/Ireland/CN-NVRL-20IRL82698/2020\|EPI_ISL_63 | ···················· | ···················· | ····················· | ···················· | ···················· | ···················· |
| R | 4145 hCoV-19/Ireland/MH-NVRL-20IRL82653/2020\|EPI_ISL_63 | ···················· | ···················· | ····················· | ···················· | ···················· | ···················· |
| R | 4146 hCoV-19/Ireland/MH-NVRL-20IRL82578/2020\|EPI_ISL_63 | ···················· | ···················· | ····················· | ···················· | ···················· | ···················· |
| R | 4147 hCoV-19/Ireland/MN-NVRL-20IRL82514/2020\|EPI_ISL_63 | ···················· | ···················· | ····················· | ···················· | ···················· | ···················· |
| R | 4148 hCoV-19/Ireland/LH-NVRL-20IRL82457/2020\|EPI_ISL_63 | ···················· | ···················· | ····················· | ···················· | ···················· | ···················· |
| R | 4149 hCoV-19/Ireland/LH-NVRL-20IRL82456/2020\|EPI_ISL_63 | ···················· | ···················· | ····················· | ···················· | ···················· | ···················· |
| R | 4150 hCoV-19/Ireland/LH-NVRL-20IRL82344/2020\|EPI_ISL_63 | ···················· | ···················· | ····················· | ···················· | ···················· | ···················· |
| R | 4151 hCoV-19/Ireland/LH-NVRL-20IRL82333/2020\|EPI_ISL_63 | ···················· | ···················· | ····················· | ···················· | ···················· | ···················· |
| R | 4152 hCoV-19/Ireland/CN-NVRL-20IRL82841/2020\|EPI_ISL_63 | ···················· | ···················· | ····················· | ···················· | ···················· | ···················· |
| R | 4153 hCoV-19/Ireland/MH-NVRL-20IRL82247/2020\|EPI_ISL_63 | ···················· | ···················· | ····················· | ···················· | ···················· | ···················· |
| R | 4154 hCoV-19/Ireland/CN-NVRL-20IRL82826/2020\|EPI_ISL_63 | ···················· | ···················· | ····················· | ···················· | ···················· | ···················· |
| R | 4155 hCoV-19/Ireland/WW-NVRL-20IRL60901/2020\|EPI_ISL_63 | ···················· | ···················· | ····················· | ···················· | ···················· | ···················· |
| R | 4156 hCoV-19/Ireland/CN-NVRL-20IRL82685/2020\|EPI_ISL_63 | ···················· | ···················· | ····················· | ···················· | ···················· | ···················· |
| R | 4157 hCoV-19/Ireland/CN-NVRL-20IRL82679/2020\|EPI_ISL_63 | ···················· | ···················· | ····················· | ···················· | ···················· | ···················· |
| R | 4158 hCoV-19/Ireland/CN-NVRL-20IRL82677/2020\|EPI_ISL_63 | ···················· | ···················· | ····················· | ···················· | ···················· | ···················· |
| R | 4159 hCoV-19/Ireland/CN-NVRL-20IRL82669/2020\|EPI_ISL_63 | ···················· | ···················· | ····················· | ···················· | ···················· | ···················· |
| R | 4160 hCoV-19/Ireland/CN-NVRL-20IRL82667/2020\|EPI_ISL_63 | ···················· | ···················· | ····················· | ···················· | ···················· | ···················· |
| R | 4161 hCoV-19/Ireland/CN-NVRL-20IRL82773/2020\|EPI_ISL_63 | ···················· | ···················· | ····················· | ···················· | ···················· | ···················· |
| R | 4162 hCoV-19/Ireland/MN-NVRL-20IRL51449/2020\|EPI_ISL_63 | ···················· | ···················· | ····················· | ···················· | ···················· | ···················· |
| R | 4163 hCoV-19/Ireland/LH-NVRL-20IRL51314/2020\|EPI_ISL_63 | ···················· | ···················· | ····················· | ···················· | ···················· | ···················· |
| R | 4164 hCoV-19/Ireland/LH-NVRL-20IRL51127/2020\|EPI_ISL_63 | ···················· | ···················· | ····················· | ···················· | ···················· | ···················· |
| R | 4165 hCoV-19/Ireland/LH-NVRL-20IRL52389/2020\|EPI_ISL_63 | ···················· | ···················· | ····················· | ···················· | ···················· | ···················· |
| R | 4166 hCoV-19/Ireland/LH-NVRL-20IRL52380/2020\|EPI_ISL_63 | ···················· | ···················· | ····················· | ···················· | ···················· | ···················· |
| R | 4167 hCoV-19/Ireland/LH-NVRL-20IRL52333/2020\|EPI_ISL_63 | ···················· | ···················· | ····················· | ···················· | ···················· | ···················· |
| R | 4168 hCoV-19/Ireland/MN-NVRL-20IRL50683/2020\|EPI_ISL_63 | ·········T·········· | ···················· | ····················· | ···················· | ···················· | ···················· |
| R | 4169 hCoV-19/Ireland/MN-NVRL-20IRL50636/2020\|EPI_ISL_63 | ···················· | ···················· | ····················· | ···················· | ···················· | ···················· |
| R | 4170 hCoV-19/Australia/VIC12494/2020\|EPI_ISL_640800\|202 | ···················· | ···················· | ····················· | ···················· | ···················· | ···················· |
| R | 4171 hCoV-19/Australia/VIC12563/2020\|EPI_ISL_640801\|202 | ···················· | ···················· | ····················· | ···················· | ···················· | ···················· |
| R | 4172 hCoV-19/Australia/VIC12898/2020\|EPI_ISL_640802\|202 | ···················· | ···················· | ····················· | ···················· | ···················· | ···················· |
| R | 4173 hCoV-19/Australia/VIC15964/2020\|EPI_ISL_640805\|202 | ···················· | ···················· | ····················· | ···················· | ···················· | ···················· |
| R | 4174 hCoV-19/Australia/VIC16480/2020\|EPI_ISL_640806\|202 | ···················· | ···················· | ····················· | ···················· | ···················· | ···················· |
| R | 4175 hCoV-19/Australia/VIC16037/2020\|EPI_ISL_640807\|202 | ···················· | ···················· | ····················· | ···················· | ···················· | ···················· |
| R | 4176 hCoV-19/Australia/VIC13275/2020\|EPI_ISL_640808\|202 | ···················· | ···················· | ····················· | ···················· | ···················· | ···················· |
| R | 4177 hCoV-19/Australia/VIC16479/2020\|EPI_ISL_640809\|202 | ···················· | ···················· | ····················· | ···················· | ···················· | ···················· |

| | | | | | | | | |
|---|---|---|---|---|---|---|---|---|
| R | 4178 hCoV-19/Australia/VIC16534/2020\|EPI_ISL_640810\|202 | ···················· | ···················· | ····················· | ···················· | ···················· | ···················· | |
| R | 4179 hCoV-19/Australia/VIC12227/2020\|EPI_ISL_640811\|202 | ···················· | ···················· | ····················· | ···················· | ···················· | ···················· | |
| R | 4180 hCoV-19/Australia/VIC13176/2020\|EPI_ISL_640812\|202 | ···················· | ···················· | ····················· | ···················· | ···················· | ···················· | |
| R | 4181 hCoV-19/Australia/VIC15987/2020\|EPI_ISL_640813\|202 | ···················· | ···················· | ····················· | ···················· | ···················· | ···················· | |
| R | 4182 hCoV-19/Australia/VIC12440/2020\|EPI_ISL_640816\|202 | ···················· | ···················· | ····················· | ···················· | ···················· | ···················· | |
| R | 4183 hCoV-19/Australia/VIC12062/2020\|EPI_ISL_640817\|202 | ···················· | ···················· | ····················· | ···················· | ···················· | ···················· | |
| R | 4184 hCoV-19/Australia/VIC16483/2020\|EPI_ISL_640819\|202 | ···················· | ···················· | ····················· | ···················· | ···················· | ···················· | |
| R | 4185 hCoV-19/Australia/VIC12907/2020\|EPI_ISL_640820\|202 | ···················· | ···················· | ····················· | ···················· | ···················· | ···················· | |
| R | 4186 hCoV-19/Australia/VIC12543/2020\|EPI_ISL_640821\|202 | ···················· | ···················· | ····················· | ···················· | ···················· | ···················· | |
| R | 4187 hCoV-19/Australia/VIC16164/2020\|EPI_ISL_640822\|202 | ···················· | ···················· | ····················· | ···················· | ···················· | ···················· | |
| R | 4188 hCoV-19/Australia/VIC15931/2020\|EPI_ISL_640823\|202 | ···················· | ···················· | ····················· | ···················· | ···················· | ···················· | |
| R | 4190 hCoV-19/Australia/VIC12589/2020\|EPI_ISL_640825\|202 | ···················· | ···················· | ····················· | ···················· | ···················· | ···················· | |
| R | 4191 hCoV-19/Australia/VIC12806/2020\|EPI_ISL_640826\|202 | ···················· | ···················· | ····················· | ···················· | ···················· | ···················· | |
| R | 4193 hCoV-19/Australia/VIC12414/2020\|EPI_ISL_640830\|202 | ···················· | ···················· | ····················· | ···················· | ···················· | ···················· | |
| R | 4195 hCoV-19/Australia/VIC16087/2020\|EPI_ISL_640832\|202 | ···················· | ···················· | ····················· | ···················· | ···················· | ···················· | |
| R | 4196 hCoV-19/Australia/VIC15946/2020\|EPI_ISL_640833\|202 | ···················· | ···················· | ····················· | ···················· | ···················· | ···················· | |
| R | 4197 hCoV-19/Australia/VIC12023/2020\|EPI_ISL_640834\|202 | ···················· | ···················· | ····················· | ···················· | ···················· | ···················· | |
| R | 4198 hCoV-19/Australia/VIC11599/2020\|EPI_ISL_640835\|202 | ···················· | ···················· | ····················· | ···················· | NNN················· | ···················· | 1 |
| R | 4199 hCoV-19/Australia/VIC12763/2020\|EPI_ISL_640837\|202 | ···················· | ···················· | ····················· | ···················· | ···················· | ···················· | |
| R | 4200 hCoV-19/Australia/VIC12017/2020\|EPI_ISL_640838\|202 | ···················· | ···················· | ····················· | ···················· | ···················· | ···················· | |
| R | 4201 hCoV-19/Australia/VIC16079/2020\|EPI_ISL_640839\|202 | ···················· | ···················· | ····················· | ···················· | ···················· | ···················· | |
| R | 4202 hCoV-19/Australia/VIC15945/2020\|EPI_ISL_640840\|202 | ···················· | ···················· | ····················· | ···················· | ···················· | ···················· | |
| R | 4203 hCoV-19/Australia/VIC16316/2020\|EPI_ISL_640841\|202 | ···················· | ···················· | ····················· | ···················· | ···················· | ···················· | |
| R | 4204 hCoV-19/Australia/VIC16664/2020\|EPI_ISL_640842\|202 | ···················· | ···················· | ····················· | ···················· | ···················· | ···················· | |
| R | 4205 hCoV-19/Australia/VIC11750/2020\|EPI_ISL_640843\|202 | ···················· | ···················· | ····················· | ···················· | ···················· | ···················· | |
| R | 4206 hCoV-19/Australia/VIC12916/2020\|EPI_ISL_640844\|202 | ···················· | ···················· | ····················· | ···················· | ···················· | ···················· | |
| R | 4207 hCoV-19/Australia/VIC12539/2020\|EPI_ISL_640845\|202 | ···················· | ···················· | ····················· | ···················· | ···················· | ···················· | |
| R | 4208 hCoV-19/Australia/VIC16120/2020\|EPI_ISL_640846\|202 | ···················· | ···················· | ····················· | ···················· | ···················· | ···················· | |
| R | 4209 hCoV-19/Australia/VIC11717/2020\|EPI_ISL_640847\|202 | ···················· | ···················· | ····················· | ···················· | ···················· | ···················· | |
| R | 4210 hCoV-19/Australia/VIC15950/2020\|EPI_ISL_640849\|202 | ···················· | ···················· | ····················· | ···················· | ···················· | ···················· | |
| R | 4211 hCoV-19/Australia/VIC12709/2020\|EPI_ISL_640850\|202 | ···················· | ···················· | ····················· | ···················· | ···················· | ···················· | |
| R | 4212 hCoV-19/Australia/VIC15990/2020\|EPI_ISL_640866\|202 | ···················· | ···················· | ····················· | ···················· | ···················· | ···················· | |
| R | 4213 hCoV-19/Australia/VIC11703/2020\|EPI_ISL_640867\|202 | ···················· | ···················· | ····················· | ···················· | ···················· | ···················· | |
| R | 4214 hCoV-19/Australia/VIC16085/2020\|EPI_ISL_640868\|202 | ···················· | ···················· | ····················· | ···················· | ···················· | ···················· | |
| R | 4215 hCoV-19/Australia/VIC11712/2020\|EPI_ISL_640869\|202 | ···················· | ···················· | ····················· | ···················· | ···················· | ···················· | |
| R | 4216 hCoV-19/Australia/VIC16300/2020\|EPI_ISL_640872\|202 | ···················· | ···················· | ····················· | ···················· | ···················· | ···················· | |
| R | 4217 hCoV-19/Australia/VIC12541/2020\|EPI_ISL_640874\|202 | ···················· | ···················· | ····················· | ···················· | ···················· | ···················· | |
| R | 4218 hCoV-19/Australia/VIC12786/2020\|EPI_ISL_640875\|202 | ···················· | ···················· | ····················· | ···················· | ···················· | ···················· | |
| R | 4220 hCoV-19/Australia/VIC12369/2020\|EPI_ISL_640877\|202 | ···················· | ···················· | ····················· | ···················· | ···················· | ···················· | |
| R | 4221 hCoV-19/Australia/VIC12815/2020\|EPI_ISL_640878\|202 | ···················· | ···················· | ····················· | ···················· | ···················· | ···················· | |
| R | 4222 hCoV-19/Australia/VIC16474/2020\|EPI_ISL_640879\|202 | ···················· | ···················· | ····················· | ···················· | ···················· | ···················· | |
| R | 4223 hCoV-19/Australia/VIC12502/2020\|EPI_ISL_640880\|202 | ···················· | ···················· | ····················· | ···················· | ···················· | ···················· | |
| R | 4224 hCoV-19/Australia/VIC16486/2020\|EPI_ISL_640881\|202 | ···················· | ···················· | ····················· | ···················· | ···················· | ···················· | |
| R | 4225 hCoV-19/Australia/VIC16365/2020\|EPI_ISL_640882\|202 | ···················· | ···················· | ····················· | ···················· | ···················· | ···················· | |
| R | 4227 hCoV-19/Australia/VIC12269/2020\|EPI_ISL_640884\|202 | ···················· | ···················· | ····················· | ···················· | ···················· | ···················· | |
| R | 4228 hCoV-19/Australia/VIC16394/2020\|EPI_ISL_640885\|202 | ···················· | ···················· | ····················· | ···················· | ···················· | ···················· | |
| R | 4229 hCoV-19/Australia/VIC12332/2020\|EPI_ISL_640886\|202 | ···················· | ···················· | ····················· | ···················· | ···················· | ···················· | |
| R | 4230 hCoV-19/Australia/VIC12725/2020\|EPI_ISL_640887\|202 | ···················· | ···················· | ····················· | ···················· | ···················· | ···················· | |
| R | 4231 hCoV-19/Australia/VIC12558/2020\|EPI_ISL_640888\|202 | ···················· | ···················· | ····················· | ···················· | ···················· | ···················· | |
| R | 4232 hCoV-19/Australia/VIC16428/2020\|EPI_ISL_640889\|202 | ···················· | ···················· | ····················· | ···················· | ···················· | ···················· | |
| R | 4233 hCoV-19/Australia/VIC11622/2020\|EPI_ISL_640890\|202 | ···················· | ···················· | ····················· | ···················· | ···················· | ···················· | |
| R | 4234 hCoV-19/Australia/VIC16449/2020\|EPI_ISL_640891\|202 | ···················· | ···················· | ····················· | ···················· | ···················· | ···················· | |
| R | 4235 hCoV-19/Australia/VIC12108/2020\|EPI_ISL_640892\|202 | ···················· | ···················· | ····················· | ···················· | ···················· | ···················· | |
| R | 4236 hCoV-19/Australia/VIC12704/2020\|EPI_ISL_640893\|202 | ···················· | ···················· | ····················· | ···················· | ···················· | ···················· | |
| R | 4237 hCoV-19/Australia/VIC16465/2020\|EPI_ISL_640894\|202 | ···················· | ···················· | ····················· | ···················· | ···················· | ···················· | |
| R | 4238 hCoV-19/Australia/VIC16375/2020\|EPI_ISL_640895\|202 | ···················· | ···················· | ····················· | ···················· | ···················· | ···················· | |
| R | 4240 hCoV-19/Australia/VIC12611/2020\|EPI_ISL_640897\|202 | ···················· | ···················· | ····················· | ···················· | ···················· | ···················· | |
| R | 4241 hCoV-19/Australia/VIC12779/2020\|EPI_ISL_640899\|202 | ···················· | ···················· | ····················· | ···················· | ···················· | ···················· | |
| R | 4242 hCoV-19/England/QEUH-A8C363/2020\|EPI_ISL_643070\|20 | ···················· | ···················· | ····················· | ···················· | ···················· | ···················· | |
| R | 4243 hCoV-19/England/QEUH-A8B4E9/2020\|EPI_ISL_643071\|20 | ···················· | ···················· | ····················· | ···················· | ···················· | ···················· | |
| R | 4244 hCoV-19/England/QEUH-A8C71C/2020\|EPI_ISL_643072\|20 | ···················· | ···················· | ····················· | ···················· | ···················· | ···················· | |
| R | 4246 hCoV-19/England/QEUH-A8C785/2020\|EPI_ISL_643074\|20 | ···················· | ···················· | ····················· | ···················· | ···················· | ···················· | |
| R | 4247 hCoV-19/England/QEUH-A8C9DA/2020\|EPI_ISL_643077\|20 | ···················· | ···················· | ····················· | ···················· | ···················· | ···················· | |
| R | 4248 hCoV-19/England/QEUH-A8BC78/2020\|EPI_ISL_643079\|20 | ···················· | ···················· | ····················· | ···················· | ···················· | ···················· | |
| R | 4249 hCoV-19/England/QEUH-A8C7B2/2020\|EPI_ISL_643080\|20 | ···················· | ···················· | ····················· | ···················· | ···················· | ···················· | |
| R | 4250 hCoV-19/England/QEUH-A8B540/2020\|EPI_ISL_643081\|20 | ···················· | ···················· | ····················· | ···················· | ···················· | ···················· | |
| R | 4251 hCoV-19/England/QEUH-A8C248/2020\|EPI_ISL_643082\|20 | ···················· | ···················· | ····················· | ···················· | ···················· | ···················· | |
| R | 4252 hCoV-19/England/QEUH-A8C99E/2020\|EPI_ISL_643083\|20 | ···················· | ···················· | ····················· | ···················· | ···················· | ···················· | |
| R | 4253 hCoV-19/England/QEUH-A8B99F/2020\|EPI_ISL_643084\|20 | ···················· | ···················· | ····················· | ···················· | ···················· | ···················· | |
| R | 4254 hCoV-19/England/QEUH-A8B56E/2020\|EPI_ISL_643085\|20 | ···················· | ···················· | ····················· | ···················· | ···················· | ···················· | |
| R | 4255 hCoV-19/England/QEUH-A8C433/2020\|EPI_ISL_643086\|20 | ···················· | ···················· | ····················· | ···················· | ···················· | ···················· | |
| R | 4256 hCoV-19/England/QEUH-A8B425/2020\|EPI_ISL_643087\|20 | ···················· | ···················· | ····················· | ···················· | ···················· | ···················· | |
| R | 4257 hCoV-19/England/QEUH-A8C257/2020\|EPI_ISL_643088\|20 | ···················· | ···················· | ····················· | ···················· | ···················· | ···················· | |
| R | 4258 hCoV-19/England/QEUH-A8BB8A/2020\|EPI_ISL_643089\|20 | ···················· | ···················· | ····················· | ···················· | ···················· | ···················· | |
| R | 4259 hCoV-19/England/QEUH-A8C0F3/2020\|EPI_ISL_643090\|20 | ···················· | ···················· | ····················· | ···················· | ···················· | ···················· | |
| R | 4260 hCoV-19/England/QEUH-A8C1D2/2020\|EPI_ISL_643091\|20 | ···················· | ·A·················· | ····················· | ···················· | ···················· | ··A················· | |
| R | 4261 hCoV-19/England/QEUH-A8B64D/2020\|EPI_ISL_643092\|20 | ···················· | ···················· | ····················· | ···················· | ···················· | ···················· | |
| R | 4262 hCoV-19/England/QEUH-A8C6B5/2020\|EPI_ISL_643093\|20 | ···················· | ···················· | ····················· | ···················· | ···················· | ···················· | |
| R | 4263 hCoV-19/England/QEUH-A8BC87/2020\|EPI_ISL_643094\|20 | ···················· | ···················· | ····················· | ···················· | ···················· | ···················· | |
| R | 4264 hCoV-19/England/QEUH-A8C837/2020\|EPI_ISL_643095\|20 | ···················· | ···················· | ····················· | ···················· | ···················· | ···················· | |

R 4265 hCoV-19/England/QEUH-A8C14B/2020|EPI_ISL_643097|20 ···················· ···················· ····················· ···················· ···················· ····················
R 4266 hCoV-19/England/QEUH-A8BE90/2020|EPI_ISL_643098|20 ···················· ···················· ····················· ···················· ···················· ····················
R 4267 hCoV-19/England/QEUH-A8BA9C/2020|EPI_ISL_643099|20 ···················· ···················· ····················· ···················· ···················· ····················
R 4268 hCoV-19/Ireland/WW-NVRL-AIIDV0425v1/2020|EPI_ISL_6 ···················· ···················· ····················· ···················· ···················· ····················
R 4269 hCoV-19/Ireland/WW-NVRL-AIIDV0351v1/2020|EPI_ISL_6 ···················· ···················· ····················· ···················· ···················· ····················
R 4270 hCoV-19/Ireland/WW-NVRL-AIIDV0351v2/2020|EPI_ISL_6 ···················· ···················· ····················· ···················· ···················· ····················
R 4271 hCoV-19/Ireland/WW-NVRL-AIIDV0063v1/2020|EPI_ISL_6 ···················· ···················· ····················· ···················· ···················· ····················
R 4272 hCoV-19/Ireland/D-NVRL-AIIDV0141v3/2020|EPI_ISL_64 ···················· ···················A ····················· ···················· ···················· ···················· 1
R 4273 hCoV-19/Ireland/D-NVRL-AIIDV0432v7/2020|EPI_ISL_64 ···················· ···················· ····················· ···················· ···················· ····················
R 4274 hCoV-19/Ireland/WW-NVRL-AIIDV0063v3/2020|EPI_ISL_6 ···················· ···················· ····················· ···················· ···················· ····················
R 4275 hCoV-19/Ireland/D-NVRL-AIIDV0346v1/2020|EPI_ISL_64 ···················· ···················· ····················· ···················· ···················· ····················
R 4276 hCoV-19/Ireland/D-NVRL-AIIDV0402v1/2020|EPI_ISL_64 ···················· ···················· ····················· ···················· ···················· ····················
R 4277 hCoV-19/Ireland/WW-NVRL-AIIDV0064v1/2020|EPI_ISL_6 ···················· ···················· ····················· ···················· ···················· ····················
R 4278 hCoV-19/Ireland/D-NVRL-AIIDV0206v1/2020|EPI_ISL_64 ···················· ···················· ····················· ···················· ···················· ····················
R 4279 hCoV-19/Ireland/WW-NVRL-AIIDV0485v1/2020|EPI_ISL_6 ···················· ···················· ····················· ···················· ···················· ····················
R 4280 hCoV-19/Ireland/WW-NVRL-AIIDV0485v2/2020|EPI_ISL_6 ···················· ···················· ····················· ···················· ···················· ····················
R 4281 hCoV-19/Ireland/WW-NVRL-AIIDV0063v4/2020|EPI_ISL_6 ···················· ···················· ····················· ···················· ···················· ····················
R 4282 hCoV-19/Ireland/D-NVRL-AIIDV0432v9/2020|EPI_ISL_64 ···················· ···················· ····················· ···················· ···················· ····················
R 4283 hCoV-19/Ireland/CE-NVRL-AIIDV1105v5/2020|EPI_ISL_6 ···················· ···················· ····················· ···················· ···················· ····················
R 4284 hCoV-19/England/ALDP-B1A026/2020|EPI_ISL_645603|20 ···················· ···················· ····················· ···················· ···················· ····················
R 4285 hCoV-19/England/ALDP-B1B9FD/2020|EPI_ISL_645604|20 ···················· ···················· ····················· ···················· ···················· ··A·················
R 4286 hCoV-19/England/ALDP-B1CD3C/2020|EPI_ISL_645605|20 ···················· ···················· ····················· ···················· ···················· ····················
R 4287 hCoV-19/England/ALDP-B1C965/2020|EPI_ISL_645606|20 ···················· ·A·················· ····················· ···················· ···················· ····················
R 4288 hCoV-19/England/ALDP-B1B580/2020|EPI_ISL_645607|20 ···················· ···················· ····················· ···················· ···················· ····················
R 4289 hCoV-19/England/ALDP-B1A114/2020|EPI_ISL_645608|20 ···················· ···················· ····················· ···················· ···················· ····················
R 4290 hCoV-19/England/ALDP-B1D0F6/2020|EPI_ISL_645609|20 ···················· ···················· ····················· ···················· ···················· ····················
R 4291 hCoV-19/England/ALDP-B1A062/2020|EPI_ISL_645610|20 ···················· ···················· ····················· ···················· ···················· ····················
R 4292 hCoV-19/England/ALDP-B1BF28/2020|EPI_ISL_645611|20 ···················· ···················· ····················· ···················· ···················· ····················
R 4293 hCoV-19/England/ALDP-B1CCC6/2020|EPI_ISL_645612|20 ···················· ···················· ····················· ···················· ···················· ····················
R 4294 hCoV-19/England/ALDP-B1BC21/2020|EPI_ISL_645613|20 ···················· ···················· ····················· ···················· ···················· ····················
R 4295 hCoV-19/England/ALDP-B1A78B/2020|EPI_ISL_645615|20 ···················· ···················· ····················· ···················· ···················· ····················
R 4296 hCoV-19/England/ALDP-B1A6F7/2020|EPI_ISL_645618|20 ···················· ···················· ····················· ···················· ···················· ····················
R 4297 hCoV-19/England/ALDP-B1ABE9/2020|EPI_ISL_645625|20 ···················· ···················· ····················· ···················· ···················· ····················
R 4298 hCoV-19/England/ALDP-B1BA09/2020|EPI_ISL_645626|20 ···················· ···················· ····················· ···················· ···················· ····················
R 4299 hCoV-19/England/ALDP-B1C3FE/2020|EPI_ISL_645627|20 ···················· ···················· ····················· ···················· ···················· ····················
R 4300 hCoV-19/England/ALDP-B1B377/2020|EPI_ISL_645628|20 ···················· ···················· ····················· ···················· ···················· ····················
R 4301 hCoV-19/England/ALDP-B1AFFC/2020|EPI_ISL_645629|20 ···················· ···················· ····················· ···················· ···················· ····················
R 4302 hCoV-19/England/ALDP-B1B0E9/2020|EPI_ISL_645630|20 ···················· ···················· ····················· ···················· ···················· ····················
R 4303 hCoV-19/England/ALDP-B1A0F9/2020|EPI_ISL_645631|20 ···················· ···················· ····················· ···················· ···················· ····················
R 4304 hCoV-19/England/ALDP-B1AF83/2020|EPI_ISL_645632|20 ···················· ···················· ····················· ···················· ···················· ····················
R 4305 hCoV-19/England/ALDP-B1B474/2020|EPI_ISL_645635|20 ···················· ···················· ····················· ···················· ···················· ····················
R 4306 hCoV-19/England/ALDP-B1C552/2020|EPI_ISL_645636|20 ···················· ···················· ····················· ···················· ···················· ····················
R 4307 hCoV-19/England/ALDP-B1A554/2020|EPI_ISL_645637|20 ···················· ···················· ····················· ···················· ···················· ····················
R 4308 hCoV-19/England/ALDP-B1B984/2020|EPI_ISL_645638|20 ···················· ···················· ····················· ···················· ···················· ····················
R 4309 hCoV-19/England/ALDP-B1AFCF/2020|EPI_ISL_645639|20 ···················· ···················· ····················· ···················· ···················· ····················
R 4310 hCoV-19/England/ALDP-B1BF46/2020|EPI_ISL_645640|20 ···················· ···················· ····················· ···················· ···················· ····················
R 4311 hCoV-19/England/ALDP-B1BACD/2020|EPI_ISL_645641|20 ···················· ···················· ····················· ···················· ···················· ····················
R 4312 hCoV-19/England/ALDP-B1CD0F/2020|EPI_ISL_645642|20 ···················· ···················· ····················· ···················· ···················· ····················
R 4313 hCoV-19/England/ALDP-B1C437/2020|EPI_ISL_645643|20 ···················· ···················· ····················· ···················· ···················· ····················
R 4314 hCoV-19/England/ALDP-B1CCB7/2020|EPI_ISL_645644|20 ···················· ···················· ····················· ···················· ···················· ····················
R 4315 hCoV-19/England/ALDP-B1A448/2020|EPI_ISL_645645|20 ···················· ···················· ····················· ···················· ···················· ····················
R 4316 hCoV-19/England/ALDP-B1B9DF/2020|EPI_ISL_645646|20 ···················· ···················· ····················· ···················· ···················· ····················
R 4317 hCoV-19/England/ALDP-B245B1/2020|EPI_ISL_646934|20 ···················· ···················· ····················· ···················· ···················· ····················
R 4318 hCoV-19/England/ALDP-B23B92/2020|EPI_ISL_646935|20 ···················· ···················· ····················· ···················· ···················· ····················
R 4319 hCoV-19/England/QEUH-B2DBF2/2020|EPI_ISL_646936|20 ···················· ···················· ····················· ···················· ···················· ····················
R 4320 hCoV-19/England/QEUH-B2DC86/2020|EPI_ISL_646937|20 ···················· ···················· ····················· ···················· ···················· ····················
R 4321 hCoV-19/England/ALDP-B243D5/2020|EPI_ISL_646938|20 ···················· ···················· ····················· ···················· ···················· ····················
R 4322 hCoV-19/England/QEUH-B2ED55/2020|EPI_ISL_646939|20 ···················· ···················· ····················· ···················· ···················· ····················
R 4323 hCoV-19/England/ALDP-B246AF/2020|EPI_ISL_646940|20 ···················· ···················· ····················· ···················· ···················· ····················
R 4324 hCoV-19/England/ALDP-B235FE/2020|EPI_ISL_646941|20 ···················· ···················· ····················· ···················· ···················· ····················
R 4325 hCoV-19/England/ALDP-B24BFB/2020|EPI_ISL_646942|20 ···················· ···················· ····················· ···················· ···················· ····················
R 4326 hCoV-19/England/ALDP-B244C3/2020|EPI_ISL_646943|20 ···················· ···················· ····················· ···················· ···················· ····················
R 4327 hCoV-19/England/ALDP-B23C71/2020|EPI_ISL_646944|20 ···················· ···················· ····················· ···················· ···················· ····················
R 4328 hCoV-19/England/ALDP-B2478E/2020|EPI_ISL_646947|20 ···················· ···················· ····················· ···················· ···················· ····················
R 4329 hCoV-19/England/ALDP-B248D6/2020|EPI_ISL_646949|20 ···················· ·A·················· ····················· ···················· ···················· ····················
R 4330 hCoV-19/England/ALDP-B2427E/2020|EPI_ISL_646957|20 ···················· ···················· ····················· ···················· ···················· ····················
R 4331 hCoV-19/England/ALDP-B24135/2020|EPI_ISL_646958|20 ···················· ···················· ····················· ···················· ···················· ····················
R 4332 hCoV-19/England/ALDP-B2360A/2020|EPI_ISL_646959|20 ···················· ···················· ····················· ···················· ···················· ····················
R 4333 hCoV-19/England/QEUH-B2EC94/2020|EPI_ISL_646960|20 ···················· ···················· ····················· ···················· ···················· ····················
R 4334 hCoV-19/England/ALDP-B24B64/2020|EPI_ISL_646961|20 ···················· ···················· ····················· ···················· ···················· ····················
R 4335 hCoV-19/England/ALDP-B23813/2020|EPI_ISL_646962|20 ···················· ···················· ····················· ···················· ···················· ····················
R 4336 hCoV-19/England/ALDP-B2451B/2020|EPI_ISL_646963|20 ···················· ···················· ····················· ···················· ·····T·············· ···············A····
R 4337 hCoV-19/England/QEUH-B2DCE0/2020|EPI_ISL_646964|20 ···················· ···················· ····················· ···················· ·····T·············· ····················
R 4338 hCoV-19/England/ALDP-B24C16/2020|EPI_ISL_646965|20 ···················· ···················· ····················· ···················· ···················· ····················
R 4339 hCoV-19/England/ALDP-B235EF/2020|EPI_ISL_646966|20 ···················· ···················· ····················· ···················· ···················· ····················
R 4340 hCoV-19/England/ALDP-B23716/2020|EPI_ISL_646968|20 ···················· ···················· ····················· ···················· ···················· ····················
R 4341 hCoV-19/England/ALDP-B2441E/2020|EPI_ISL_646969|20 ···················· ···················· ····················· ···················· ···················· ····················
R 4342 hCoV-19/England/ALDP-B24180/2020|EPI_ISL_646971|20 ···················· ···················· ····················· ···················· ···················· ····················
R 4343 hCoV-19/England/ALDP-B2740C/2020|EPI_ISL_646972|20 ···················· ···················· ····················· ···················· ···················· ····················
R 4344 hCoV-19/England/ALDP-B2393E/2020|EPI_ISL_646973|20 ···················· ···················· ····················· ···················· ···················· ····················

| | | | | | | | | |
|---|---|---|---|---|---|---|---|---|
| R | 4345 | hCoV-19/England/QEUH-B2DC4A/2020\|EPI_ISL_646974\|20 | ···················· | ···················· | ···················· | ···················· | ···················· | ···················· |
| R | 4346 | hCoV-19/England/QEUH-B2DA31/2020\|EPI_ISL_646975\|20 | ···················· | ·A··G··············· | ···················· | ···················· | ···················· | ···················· |
| R | 4347 | hCoV-19/England/QEUH-B2D73A/2020\|EPI_ISL_646976\|20 | ···················· | ···················· | ···················· | ···················· | ···················· | ···················· |
| R | 4348 | hCoV-19/England/QEUH-B2DC77/2020\|EPI_ISL_646977\|20 | ···················· | ···················· | ···················· | ···················· | ···················· | ···················· |
| R | 4349 | hCoV-19/cat/USA/TX-TAMU-013/2020\|EPI_ISL_699506\|20 | ···················· | ·A·················· | ···················· | ···················· | ···················· | ···················· |
| R | 4350 | hCoV-19/cat/USA/TX-TAMU-057/2020\|EPI_ISL_699507\|20 | ···················· | ···················· | ···················· | ···················· | ···················· | ···················· |
| R | 4351 | hCoV-19/dog/USA/TX-TAMU-077/2020\|EPI_ISL_699508\|20 | ···················· | ···················· | ···················· | ···················· | ···················· | ···················· |
| R | 4352 | hCoV-19/cat/USA/TX-TAMU-078/2020\|EPI_ISL_699509\|20 | ···················· | ···················· | ···················· | ···················· | ···················· | ···················· |
| R | 4353 | hCoV-19/Australia/QLD1124/2020\|EPI_ISL_639741\|2020 | ···················· | ···················· | ···················· | ···················· | ···················· | ···················· |
| R | 4354 | hCoV-19/Australia/QLD1125/2020\|EPI_ISL_639742\|2020 | ···················· | ···················· | ···················· | ···················· | ···················· | ···················· |
| R | 4355 | hCoV-19/Australia/QLD1126/2020\|EPI_ISL_639743\|2020 | ···················· | ···················· | ···················· | ···················· | ···················· | ···················· |
| R | 4356 | hCoV-19/Australia/QLD1127/2020\|EPI_ISL_639744\|2020 | ···················· | ···················· | ···················· | ···················· | ···················· | ···················· |
| R | 4357 | hCoV-19/Australia/QLD1128/2020\|EPI_ISL_639745\|2020 | ···················· | ···················· | ···················· | ···················· | ···················· | ···················· |
| R | 4358 | hCoV-19/Australia/QLD1129/2020\|EPI_ISL_639746\|2020 | ···················· | ···················· | ···················· | ···················· | ···················· | ···················· |
| R | 4359 | hCoV-19/Australia/QLD1130/2020\|EPI_ISL_639747\|2020 | ···················· | ···················· | ···················· | ···················· | ···················· | ···················· |
| R | 4360 | hCoV-19/Australia/QLD1131/2020\|EPI_ISL_639748\|2020 | ···················· | ···················· | ···················· | ···················· | ···················· | ···················· |
| R | 4361 | hCoV-19/Australia/QLD1116/2020\|EPI_ISL_639749\|2020 | ···················· | ···················· | ···················· | ···················· | ···················· | ···················· |
| R | 4362 | hCoV-19/Australia/QLD1209/2020\|EPI_ISL_639800\|2020 | ···················· | ···················· | ···················· | ···················· | ···················· | ···················· |
| R | 4363 | hCoV-19/Australia/QLD1210/2020\|EPI_ISL_639801\|2020 | ···················· | ···················· | ···················· | ···················· | ···················· | ···················· |
| R | 4364 | hCoV-19/Australia/QLD1211/2020\|EPI_ISL_639802\|2020 | ···················· | ···················· | ···················· | ···················· | ···················· | ···················· |
| R | 4365 | hCoV-19/Australia/QLD1212/2020\|EPI_ISL_639803\|2020 | ···················· | ···················· | ···················· | ···················· | ···················· | ···················· |
| R | 4366 | hCoV-19/Australia/QLD1213/2020\|EPI_ISL_639804\|2020 | ···················· | ···················· | ···················· | ···················· | ···················· | ···················· |
| R | 4367 | hCoV-19/Australia/QLD1214/2020\|EPI_ISL_639805\|2020 | ···················· | ···················· | ···················· | ···················· | ···················· | ···················· |
| R | 4368 | hCoV-19/Australia/VIC12617/2020\|EPI_ISL_640730\|202 | ···················· | ···················· | ···················· | ···················· | ···················· | ···················· |
| R | 4369 | hCoV-19/Australia/VIC16685/2020\|EPI_ISL_640731\|202 | ···················· | ···················· | ···················· | ···················· | ···················· | ···················· |
| R | 4370 | hCoV-19/Australia/VIC12376/2020\|EPI_ISL_640733\|202 | ···················· | ···················· | ···················· | ···················· | ···················· | ···················· |
| R | 4371 | hCoV-19/Australia/VIC16305/2020\|EPI_ISL_640734\|202 | ···················· | ···················· | ···················· | ···················· | ···················· | ···················· |
| R | 4372 | hCoV-19/Australia/VIC16496/2020\|EPI_ISL_640735\|202 | ···················· | ···················· | ···················· | ···················· | ···················· | ···················· |
| R | 4373 | hCoV-19/Australia/VIC16537/2020\|EPI_ISL_640736\|202 | ···················· | ···················· | ···················· | ···················· | ···················· | ···················· |
| R | 4374 | hCoV-19/Australia/VIC16105/2020\|EPI_ISL_640737\|202 | ···················· | ···················· | ···················· | ···················· | ···················· | ···················· |
| R | 4375 | hCoV-19/Australia/VIC12012/2020\|EPI_ISL_640743\|202 | ···················· | ···················· | ···················· | ···················· | ···················· | ···················· |
| R | 4377 | hCoV-19/Australia/VIC13102/2020\|EPI_ISL_640746\|202 | ···················· | ···················· | ···················· | ···················· | ···················· | ···················· |
| R | 4378 | hCoV-19/Australia/VIC16694/2020\|EPI_ISL_640747\|202 | ···················· | ···················· | ···················· | ···················· | ···················· | ···················· |
| R | 4379 | hCoV-19/Australia/VIC16177/2020\|EPI_ISL_640749\|202 | ···················· | ···················· | ···················· | ···················· | ···················· | ···················· |
| R | 4380 | hCoV-19/Australia/VIC12234/2020\|EPI_ISL_640750\|202 | ···················· | ···················· | ···················· | ···················· | ···················· | ···················· |
| R | 4381 | hCoV-19/England/MILK-B05F03/2020\|EPI_ISL_643000\|20 | ···················· | ···················· | ···················· | ···················· | ···················· | ···················· |
| R | 4382 | hCoV-19/England/MILK-B02C5A/2020\|EPI_ISL_643001\|20 | ···················· | ···················· | ···················· | ···················· | ···················· | ···················· |
| R | 4383 | hCoV-19/England/MILK-B01FBC/2020\|EPI_ISL_643002\|20 | ···················· | ···················· | ···················· | ···················· | ···················· | ···················· |
| R | 4384 | hCoV-19/England/MILK-B026D4/2020\|EPI_ISL_643003\|20 | ···················· | ···················· | ···················· | ···················· | ···················· | ···················· |
| R | 4385 | hCoV-19/England/MILK-B02847/2020\|EPI_ISL_643004\|20 | ···················· | ···················· | ···················· | ···················· | ···················· | ···················· |
| R | 4386 | hCoV-19/England/MILK-B02B6C/2020\|EPI_ISL_643005\|20 | ···················· | ·A·················· | ···················· | ···················· | ···················· | ··A················· |
| R | 4387 | hCoV-19/England/MILK-B020B8/2020\|EPI_ISL_643007\|20 | ···················· | ·A·················· | ···················· | ···················· | ···················· | ··A················· |
| R | 4388 | hCoV-19/England/MILK-B05F21/2020\|EPI_ISL_643009\|20 | ···················· | ···················· | ···················· | ···················· | ···················· | ···················· |
| R | 4389 | hCoV-19/England/MILK-B02601/2020\|EPI_ISL_643010\|20 | ···················· | ···················· | ···················· | ···················· | ···················· | ···················· |
| R | 4390 | hCoV-19/England/MILK-B02D84/2020\|EPI_ISL_643011\|20 | ···················· | ···················· | ···················· | ···················· | ···················· | ···················· |
| R | 4391 | hCoV-19/England/MILK-B0612A/2020\|EPI_ISL_643012\|20 | ···················· | ···················· | ···················· | ···················· | ···················· | ···················· |
| R | 4392 | hCoV-19/England/MILK-B020D6/2020\|EPI_ISL_643013\|20 | ···················· | ···················· | ···················· | ···················· | ···················· | ···················· |
| R | 4393 | hCoV-19/England/MILK-B023A0/2020\|EPI_ISL_643014\|20 | ···················· | ···················· | ···················· | ···················· | ···················· | ···················· |
| R | 4394 | hCoV-19/England/MILK-B06069/2020\|EPI_ISL_643015\|20 | ···················· | ···················· | ···················· | ···················· | ······T············· | ···················· |
| R | 4395 | hCoV-19/England/MILK-B06209/2020\|EPI_ISL_643016\|20 | ···················· | ···················· | ···················· | ···················· | ···················· | ···················· |
| R | 4396 | hCoV-19/England/MILK-B05FA9/2020\|EPI_ISL_643017\|20 | ···················· | ···················· | ···················· | ···················· | ···················· | ···················· |
| R | 4397 | hCoV-19/England/MILK-B02E18/2020\|EPI_ISL_643018\|20 | ···················· | ···················· | ···················· | ···················· | ···················· | ···················· |
| R | 4398 | hCoV-19/England/MILK-B06281/2020\|EPI_ISL_643019\|20 | ···················· | ···················· | ···················· | ···················· | ···················· | ···················· |
| R | 4399 | hCoV-19/England/MILK-B021D3/2020\|EPI_ISL_643021\|20 | ···················· | ·A·················· | ···················· | ···················· | ···················· | ··A················· |
| R | 4400 | hCoV-19/England/MILK-B0262F/2020\|EPI_ISL_643022\|20 | ···················· | ···················· | ···················· | ···················· | ···················· | ···················· |
| R | 4401 | hCoV-19/England/MILK-B06351/2020\|EPI_ISL_643023\|20 | ···················· | ···················· | ···················· | ···················· | ···················· | ···················· |
| R | 4402 | hCoV-19/England/MILK-B061DF/2020\|EPI_ISL_643024\|20 | ···················· | ···················· | ···················· | ···················· | ···················· | ···················· |
| R | 4403 | hCoV-19/England/MILK-B05F7C/2020\|EPI_ISL_643025\|20 | ···················· | ···················· | ···················· | ···················· | ···················· | ···················· |
| R | 4404 | hCoV-19/England/MILK-B02AE7/2020\|EPI_ISL_643026\|20 | ···················· | ···················· | ···················· | ···················· | ···················· | ···················· |
| R | 4405 | hCoV-19/England/MILK-B063AC/2020\|EPI_ISL_643027\|20 | ···················· | ···················· | ···················· | ···················· | ···················· | ···················· |
| R | 4406 | hCoV-19/England/MILK-B02A7E/2020\|EPI_ISL_643028\|20 | ···················· | ···················· | ···················· | ···················· | ······T············· | ···················· |
| R | 4407 | hCoV-19/England/MILK-B02D0C/2020\|EPI_ISL_643030\|20 | ···················· | ···················· | ···················· | ···················· | ···················· | ···················· |
| R | 4408 | hCoV-19/England/MILK-B05ECA/2020\|EPI_ISL_643031\|20 | ···················· | ···················· | ···················· | ···················· | ···················· | ···················· |
| R | 4409 | hCoV-19/England/MILK-B06184/2020\|EPI_ISL_643032\|20 | ···················· | ···················· | ···················· | ···················· | ···················· | ···················· |
| R | 4410 | hCoV-19/England/MILK-B01F9E/2020\|EPI_ISL_643033\|20 | ···················· | ···················· | ···················· | ···················· | ···················· | ···················· |
| R | 4411 | hCoV-19/England/MILK-B026F2/2020\|EPI_ISL_643034\|20 | ···················· | ···················· | ···················· | ···················· | ···················· | ···················· |
| R | 4412 | hCoV-19/England/MILK-B02856/2020\|EPI_ISL_643035\|20 | ···················· | ···················· | ···················· | ···················· | ···················· | ···················· |
| R | 4413 | hCoV-19/England/MILK-B01D58/2020\|EPI_ISL_643036\|20 | ···················· | ···················· | ···················· | ···················· | ···················· | ···················· |
| R | 4414 | hCoV-19/England/MILK-B01D67/2020\|EPI_ISL_643037\|20 | ···················· | ·A·················· | ···················· | ···················· | ···················· | ··A················· |
| R | 4415 | hCoV-19/England/MILK-B02829/2020\|EPI_ISL_643038\|20 | ···················· | ···················· | ···················· | ···················· | ···················· | ···················· |
| R | 4416 | hCoV-19/England/MILK-B01E55/2020\|EPI_ISL_643039\|20 | ···················· | ···················· | ···················· | ···················· | ···················· | ···················· |
| R | 4417 | hCoV-19/England/MILK-B01FDA/2020\|EPI_ISL_643040\|20 | ···················· | ···················· | ···················· | ···················· | ···················· | ···················· |
| R | 4418 | hCoV-19/England/MILK-B01F07/2020\|EPI_ISL_643041\|20 | ···················· | ···················· | ···················· | ···················· | ···················· | ···················· |
| R | 4419 | hCoV-19/England/MILK-B024CB/2020\|EPI_ISL_643042\|20 | ···················· | ···················· | ···················· | ···················· | ···················· | ···················· |
| R | 4420 | hCoV-19/England/MILK-B0258C/2020\|EPI_ISL_643043\|20 | ···················· | ···················· | ···················· | ···················· | ···················· | ···················· |
| R | 4421 | hCoV-19/England/QEUH-A8BCA5/2020\|EPI_ISL_643044\|20 | ···················· | ···················· | ···················· | ···················· | ···················· | ···················· |
| R | 4422 | hCoV-19/England/QEUH-A8B328/2020\|EPI_ISL_643045\|20 | ···················· | ···················· | ···················· | ···················· | ···················· | ···················· |
| R | 4423 | hCoV-19/England/QEUH-A8B49E/2020\|EPI_ISL_643046\|20 | ···················· | ···················· | ···················· | ···················· | ···················· | ···················· |
| R | 4424 | hCoV-19/England/QEUH-A8B8ED/2020\|EPI_ISL_643047\|20 | ···················· | ···················· | ···················· | ···················· | ···················· | ···················· |
| R | 4425 | hCoV-19/England/QEUH-A8C70D/2020\|EPI_ISL_643048\|20 | ···················· | ···················· | ···················· | ···················· | ···················· | ···················· |

| | | | | | | | | | |
|---|---|---|---|---|---|---|---|---|---|
| R | 4426 | hCoV-19/England/QEUH-A8BDFD/2020\|EPI_ISL_643049\|20 | .................... | .................... | .................... | .................... | .................... | .................... | |
| R | 4427 | hCoV-19/England/QEUH-A8C187/2020\|EPI_ISL_643050\|20 | .................... | .................... | .................... | .................... | .................... | .................... | |
| R | 4428 | hCoV-19/England/QEUH-A8BE45/2020\|EPI_ISL_643051\|20 | .................... | .................... | .................... | .................... | .................... | .................... | |
| R | 4429 | hCoV-19/England/QEUH-A8C0C6/2020\|EPI_ISL_643052\|20 | .................... | .................... | .................... | .................... | .................... | .................... | |
| R | 4430 | hCoV-19/England/QEUH-A8C15A/2020\|EPI_ISL_643053\|20 | .................... | .................... | .................... | .................... | .................... | .................... | |
| R | 4431 | hCoV-19/England/QEUH-A8BA6F/2020\|EPI_ISL_643054\|20 | .................... | .................... | .................... | .................... | .................... | .................... | |
| R | 4432 | hCoV-19/England/QEUH-A8B5F5/2020\|EPI_ISL_643055\|20 | .................... | .................... | .................... | .................... | .................... | .................... | |
| R | 4433 | hCoV-19/England/QEUH-A8B5B9/2020\|EPI_ISL_643057\|20 | .................... | .................... | .................... | .................... | ......T............. | .................... | |
| R | 4434 | hCoV-19/England/QEUH-A8B5D7/2020\|EPI_ISL_643058\|20 | .................... | .................... | .................... | .................... | .................... | .................... | |
| R | 4435 | hCoV-19/England/QEUH-A8BFD9/2020\|EPI_ISL_643059\|20 | .................... | .................... | .................... | .................... | .................... | .................... | |
| R | 4436 | hCoV-19/England/QEUH-A8BD75/2020\|EPI_ISL_643060\|20 | .................... | .................... | .................... | .................... | .................... | .................... | |
| R | 4437 | hCoV-19/England/QEUH-A8B7C2/2020\|EPI_ISL_643061\|20 | .................... | .................... | .................... | .................... | .................... | .................... | |
| R | 4438 | hCoV-19/England/QEUH-A8BD57/2020\|EPI_ISL_643062\|20 | .................... | .................... | .................... | .................... | .................... | .................... | |
| R | 4439 | hCoV-19/England/QEUH-A8C6E2/2020\|EPI_ISL_643063\|20 | .................... | .................... | .................... | .................... | .................... | .................... | |
| R | 4440 | hCoV-19/England/QEUH-A8B892/2020\|EPI_ISL_643065\|20 | .................... | .................... | .................... | .................... | .................... | .................... | |
| R | 4441 | hCoV-19/England/QEUH-A8BF15/2020\|EPI_ISL_643066\|20 | ..T................. | .................... | .................... | .................... | .................... | .................... | |
| R | 4442 | hCoV-19/England/QEUH-A8CA04/2020\|EPI_ISL_643067\|20 | .................... | .................... | .................... | .................... | .................... | .................... | |
| R | 4443 | hCoV-19/England/QEUH-A8C442/2020\|EPI_ISL_643068\|20 | .................... | .................... | .................... | .................... | .................... | .................... | |
| R | 4444 | hCoV-19/England/QEUH-A8B4F8/2020\|EPI_ISL_643069\|20 | .................... | .................... | .................... | .................... | .................... | .................... | |
| R | 4445 | hCoV-19/Ireland/D-NVRL-AIIDV0071v1/2020\|EPI_ISL_64 | ..................T. | .................... | .................... | .................... | .................... | .................... | 1 |
| R | 4446 | hCoV-19/Ireland/CE-NVRL-AIIDV1105v6/2020\|EPI_ISL_6 | .................... | .................... | .................... | .................... | .................... | .................... | |
| R | 4447 | hCoV-19/Ireland/D-NVRL-AIIDV0084v1/2020\|EPI_ISL_64 | .................... | .................... | .................... | .................... | .................... | .................... | |
| R | 4448 | hCoV-19/Ireland/D-NVRL-AIIDV0019v1/2020\|EPI_ISL_64 | .................... | .................... | .................... | .................... | .................... | .................... | |
| R | 4449 | hCoV-19/Ireland/WW-NVRL-AIIDV0018v1/2020\|EPI_ISL_6 | .................... | .................... | .................... | .................... | .................... | .................... | |
| R | 4450 | hCoV-19/Ireland/D-NVRL-AIIDV0002v1/2020\|EPI_ISL_64 | .................... | .................... | .................... | .................... | .................... | .................... | |
| R | 4451 | hCoV-19/Ireland/D-NVRL-AIIDV0102v2/2020\|EPI_ISL_64 | .................... | .................N.. | .................... | .................... | .................... | .................... | 1 |
| R | 4452 | hCoV-19/Ireland/D-NVRL-AIIDV0363v1/2020\|EPI_ISL_64 | .................... | .................... | .................... | .................... | .................... | .................... | |
| R | 4453 | hCoV-19/Ireland/D-NVRL-AIIDV0432v10/2020\|EPI_ISL_6 | .................... | .................... | .................... | .................... | .................... | .................... | |
| R | 4454 | hCoV-19/Ireland/D-NVRL-AIIDV0205v1/2020\|EPI_ISL_64 | .................... | ...................N | .................... | .................... | .................... | .................... | 1 |
| R | 4455 | hCoV-19/Ireland/D-NVRL-AIIDV0030v1/2020\|EPI_ISL_64 | .................... | .................... | .................... | .................... | .................... | .................... | |
| R | 4456 | hCoV-19/Ireland/WW-NVRL-AIIDV0063v5/2020\|EPI_ISL_6 | .................... | .................... | .................... | .................... | .................... | .................... | |
| R | 4457 | hCoV-19/Ireland/D-NVRL-AIIDV0072v4/2020\|EPI_ISL_64 | .................... | .................... | .................... | .................... | .................... | .................... | |
| R | 4458 | hCoV-19/Ireland/D-NVRL-AIIDV0380v1/2020\|EPI_ISL_64 | .................... | .................... | .................... | .................... | .................... | .................... | |
| R | 4459 | hCoV-19/Ireland/WW-NVRL-AIIDV0379v1/2020\|EPI_ISL_6 | .................... | .................... | .................... | .................... | .................... | .................... | |
| R | 4460 | hCoV-19/Ireland/D-NVRL-AIIDV0098v1/2020\|EPI_ISL_64 | .................... | .................... | .................... | .................... | .................... | .................... | |
| R | 4461 | hCoV-19/Ireland/D-NVRL-AIIDV0076v1/2020\|EPI_ISL_64 | .................... | .................... | .................... | .................... | .................... | .................... | |
| R | 4462 | hCoV-19/Ireland/D-NVRL-AIIDV0091v1/2020\|EPI_ISL_64 | .................... | .................... | .................... | .................... | .................... | .................... | |
| R | 4463 | hCoV-19/Ireland/WW-NVRL-AIIDV0063v6/2020\|EPI_ISL_6 | .................... | .................... | .................... | .................... | .................... | .................... | |
| R | 4464 | hCoV-19/Ireland/D-NVRL-AIIDV0053v2/2020\|EPI_ISL_64 | .................... | .................... | .................... | .................... | .................... | .................... | |
| R | 4465 | hCoV-19/Ireland/D-NVRL-AIIDV0361v2/2020\|EPI_ISL_64 | .................... | .................... | .................... | .................... | .................... | .................... | |
| R | 4466 | hCoV-19/Ireland/WW-NVRL-AIIDV0379v3/2020\|EPI_ISL_6 | .................... | .................... | .................... | .................... | .................... | .................... | |
| R | 4467 | hCoV-19/Ireland/CE-NVRL-AIIDV1105v7/2020\|EPI_ISL_6 | .................... | .................... | .................... | .................... | .................... | .................... | |
| R | 4468 | hCoV-19/Ireland/D-NVRL-AIIDV0090v2/2020\|EPI_ISL_64 | .................... | .................... | .................... | .................... | .................... | .................... | |
| R | 4469 | hCoV-19/Ireland/D-NVRL-AIIDV0022v1/2020\|EPI_ISL_64 | .................... | .................... | .................... | .................... | .................... | .................... | |
| R | 4470 | hCoV-19/Ireland/D-NVRL-AIIDV0328v2/2020\|EPI_ISL_64 | .................... | .................... | .................... | .................... | .................... | .................... | |
| R | 4471 | hCoV-19/Ireland/D-NVRL-AIIDV1114v1/2020\|EPI_ISL_64 | .................... | .................... | .................... | .................... | .................... | .................... | |
| R | 4472 | hCoV-19/Ireland/WW-NVRL-AIIDV0010v1/2020\|EPI_ISL_6 | .................... | .................... | .................... | .................... | .................... | .................... | |
| R | 4473 | hCoV-19/Ireland/D-NVRL-AIIDV0380v3/2020\|EPI_ISL_64 | .................... | .................... | .................... | .................... | .................... | .................... | |
| R | 4474 | hCoV-19/Ireland/WW-NVRL-AIIDV0379v4/2020\|EPI_ISL_6 | .................... | .................... | .................... | .................... | .................... | .................... | |
| R | 4475 | hCoV-19/USA/MI-MDHHS-SC22196/2020\|EPI_ISL_644364\|2 | .................... | .................... | .................... | .................... | .................... | .................... | |
| R | 4476 | hCoV-19/USA/MI-MDHHS-SC22197/2020\|EPI_ISL_644365\|2 | .................... | .................... | .................... | .................... | .................... | .................... | |
| R | 4477 | hCoV-19/USA/MI-MDHHS-SC22198/2020\|EPI_ISL_644366\|2 | .................... | .A.................. | .................... | .................... | .................... | .................... | |
| R | 4478 | hCoV-19/USA/MI-MDHHS-SC22199/2020\|EPI_ISL_644367\|2 | .................... | .............A...... | .................... | .................... | .................... | .................... | |
| R | 4479 | hCoV-19/USA/MI-MDHHS-SC22200/2020\|EPI_ISL_644368\|2 | .................... | .A.................. | .................... | .................... | .................... | .................... | |
| R | 4480 | hCoV-19/USA/MI-MDHHS-SC22201/2020\|EPI_ISL_644369\|2 | .................... | .............A...... | .................... | .................... | .................... | .................... | |
| R | 4481 | hCoV-19/USA/MI-MDHHS-SC22202/2020\|EPI_ISL_644370\|2 | .................... | .A.................. | .................... | .................... | .................... | .................... | |
| R | 4482 | hCoV-19/USA/MI-MDHHS-SC22203/2020\|EPI_ISL_644371\|2 | .................... | .................... | .................... | .................... | .................... | .................... | |
| R | 4483 | hCoV-19/USA/MI-MDHHS-SC22204/2020\|EPI_ISL_644372\|2 | .................... | .............A...... | .................... | .................... | .................... | .................... | |
| R | 4484 | hCoV-19/USA/MI-MDHHS-SC22205/2020\|EPI_ISL_644373\|2 | .................... | .................... | .................... | .................... | .................... | .................... | |
| R | 4485 | hCoV-19/USA/MI-MDHHS-SC22206/2020\|EPI_ISL_644374\|2 | .................... | .A.................. | .................... | .................... | .................... | .................... | |
| R | 4486 | hCoV-19/France/PAC-MEPHI-1752/2020\|EPI_ISL_644392\| | .................... | .................... | .................... | .................... | .................... | .................... | |
| R | 4487 | hCoV-19/France/PAC-MEPHI-1665/2020\|EPI_ISL_644393\| | .................... | .................... | .................... | .................... | .................... | .................... | |
| R | 4488 | hCoV-19/France/PAC-MEPHI-1666/2020\|EPI_ISL_644394\| | .................... | .................... | .................... | .................... | .................... | .................... | |
| R | 4489 | hCoV-19/France/PAC-MEPHI-1672/2020\|EPI_ISL_644395\| | .................... | .................... | .................... | .................... | .................... | .................... | |
| R | 4490 | hCoV-19/France/PAC-MEPHI-1756/2020\|EPI_ISL_644396\| | .................... | .................... | .................... | .................... | .................... | .................... | |
| R | 4491 | hCoV-19/France/PAC-MEPHI-2323/2020\|EPI_ISL_644397\| | .................... | .................... | .................... | .................... | .................... | .................... | |
| R | 4492 | hCoV-19/France/PAC-MEPHI-2361/2020\|EPI_ISL_644399\| | .................... | .................... | .................... | .................... | .................... | .................... | |
| R | 4493 | hCoV-19/England/ALDP-B1B3E0/2020\|EPI_ISL_645600\|20 | .................... | .................... | .................... | .................... | .................... | .................... | |
| R | 4494 | hCoV-19/England/ALDP-B1B939/2020\|EPI_ISL_645601\|20 | .................... | .................... | .................... | .................... | .................... | .................... | |
| R | 4495 | hCoV-19/England/ALDP-B1BA27/2020\|EPI_ISL_645602\|20 | .................... | .................... | .................... | .................... | .................... | ..A................. | |
| R | 4496 | hCoV-19/England/ALDP-B1B4FC/2020\|EPI_ISL_645614\|20 | .................... | .................... | .................... | .................... | .................... | .................... | |
| R | 4497 | hCoV-19/England/ALDP-B1B17D/2020\|EPI_ISL_645616\|20 | .................... | .................... | .................... | .................... | .................... | .................... | |
| R | 4498 | hCoV-19/England/ALDP-B1B034/2020\|EPI_ISL_645617\|20 | .................... | .................... | .................... | .................... | .................... | .................... | |
| R | 4499 | hCoV-19/England/ALDP-B1B553/2020\|EPI_ISL_645620\|20 | .................... | .................... | .................... | .................... | .................... | .................... | |
| R | 4500 | hCoV-19/England/ALDP-B1B3D1/2020\|EPI_ISL_645621\|20 | .................... | .................... | .................... | .................... | .................... | .................... | |
| R | 4501 | hCoV-19/England/ALDP-B1A1AB/2020\|EPI_ISL_645623\|20 | .................... | .................... | .................... | .................... | .................... | .................... | |
| R | 4502 | hCoV-19/England/ALDP-B1AF56/2020\|EPI_ISL_645624\|20 | .................... | .................... | .................... | .................... | .................... | .................... | |
| R | 4503 | hCoV-19/England/ALDP-B1B0BC/2020\|EPI_ISL_645647\|20 | .................... | .................... | .................... | .................... | .................... | .................... | |
| R | 4504 | hCoV-19/England/ALDP-B1B15F/2020\|EPI_ISL_645648\|20 | .................... | .................... | .................... | .................... | .................... | .................... | |
| R | 4505 | hCoV-19/England/ALDP-B1B526/2020\|EPI_ISL_645649\|20 | .................... | .................... | .................... | .................... | .................... | .................... | |

| | | | | | | | |
|---|---|---|---|---|---|---|---|
| R | 4506 hCoV-19/England/ALDP-B1BBCA/2020\|EPI_ISL_645650\|20 | ···················· | ···················· | ···················· | ···················· | ···················· | ···················· |
| R | 4507 hCoV-19/England/ALDP-B1BAEB/2020\|EPI_ISL_645651\|20 | ···················· | ···················· | ···················· | ···················· | ···················· | ···················· |
| R | 4508 hCoV-19/England/ALDP-B1CE39/2020\|EPI_ISL_645652\|20 | ···················· | ···················· | ···················· | ···················· | ···················· | ···················· |
| R | 4509 hCoV-19/England/ALDP-B1CDE1/2020\|EPI_ISL_645653\|20 | ···················· | ·A·················· | ···················· | ···················· | ···················· | ···················· |
| R | 4510 hCoV-19/England/ALDP-B1C33A/2020\|EPI_ISL_645655\|20 | ···················· | ···················· | ···················· | ···················· | ···················· | ···················· |
| R | 4511 hCoV-19/England/ALDP-B1BFA0/2020\|EPI_ISL_645656\|20 | ···················· | ···················· | ···················· | ···················· | ···················· | ···················· |
| R | 4512 hCoV-19/England/ALDP-B1B91B/2020\|EPI_ISL_645657\|20 | ···················· | ···················· | ···················· | ···················· | ······T············· | ···················· |
| R | 4513 hCoV-19/England/ALDP-B1C525/2020\|EPI_ISL_645658\|20 | ···················· | ···················· | ···················· | ···················· | ···················· | ···················· |
| R | 4514 hCoV-19/England/ALDP-B1A642/2020\|EPI_ISL_645660\|20 | ···················· | ···················· | ···················· | ···················· | ···················· | ···················· |
| R | 4515 hCoV-19/England/ALDP-B1BC03/2020\|EPI_ISL_645661\|20 | ···················· | ···················· | ···················· | ···················· | ···················· | ···················· |
| R | 4516 hCoV-19/England/ALDP-B1C5AD/2020\|EPI_ISL_645662\|20 | ···················· | ···················· | ···················· | ···················· | ···················· | ···················· |
| R | 4517 hCoV-19/England/ALDP-B1ABCB/2020\|EPI_ISL_645663\|20 | ···················· | ···················· | ···················· | ···················· | ···················· | ···················· |
| R | 4518 hCoV-19/England/ALDP-B1B92A/2020\|EPI_ISL_645664\|20 | ···················· | ···················· | ···················· | ···················· | ···················· | ···················· |
| R | 4519 hCoV-19/England/ALDP-B1B26B/2020\|EPI_ISL_645665\|20 | ···················· | ···················· | ···················· | ···················· | ···················· | ···················· |
| R | 4520 hCoV-19/England/ALDP-B1C3B2/2020\|EPI_ISL_645666\|20 | ···················· | ·········T·········· | ···················· | ···················· | ···················· | ···················· |
| R | 4521 hCoV-19/England/ALDP-B1B465/2020\|EPI_ISL_645667\|20 | ···················· | ···················· | ···················· | ···················· | ···················· | ···················· |
| R | 4522 hCoV-19/England/ALDP-B1CD1E/2020\|EPI_ISL_645668\|20 | ···················· | ···················· | ···················· | ···················· | ···················· | ···················· |
| R | 4523 hCoV-19/England/ALDP-B1C4BF/2020\|EPI_ISL_645669\|20 | ···················· | ···················· | ···················· | ···················· | ···················· | ···················· |
| R | 4524 hCoV-19/England/ALDP-B1BAFA/2020\|EPI_ISL_645670\|20 | ···················· | ···················· | ···················· | ···················· | ···················· | ···················· |
| R | 4525 hCoV-19/England/ALDP-B1B2E3/2020\|EPI_ISL_645673\|20 | ···················· | ···················· | ···················· | ···················· | ···················· | ···················· |
| R | 4526 hCoV-19/England/ALDP-B1C5BC/2020\|EPI_ISL_645674\|20 | ···················· | ···················· | ···················· | ···················· | ···················· | ···················· |
| R | 4527 hCoV-19/England/ALDP-B1BF19/2020\|EPI_ISL_645675\|20 | ···················· | ···················· | ···················· | ···················· | ···················· | ···················· |
| R | 4528 hCoV-19/England/ALDP-B1CCE4/2020\|EPI_ISL_645676\|20 | ···················· | ···················· | ···················· | ···················· | ···················· | ···················· |
| R | 4529 hCoV-19/England/ALDP-B1CD87/2020\|EPI_ISL_645677\|20 | ···················· | ···················· | ···················· | ···················· | ···················· | ···················· |
| R | 4530 hCoV-19/England/ALDP-B1B19B/2020\|EPI_ISL_645678\|20 | ···················· | ···················· | ···················· | ···················· | ···················· | ···················· |
| R | 4531 hCoV-19/England/ALDP-B1B605/2020\|EPI_ISL_645679\|20 | ···················· | ···················· | ···················· | ···················· | ···················· | ···················· |
| R | 4532 hCoV-19/England/ALDP-B1B061/2020\|EPI_ISL_645680\|20 | ···················· | ···················· | ···················· | ···················· | ···················· | ···················· |
| R | 4533 hCoV-19/England/ALDP-B1AFA1/2020\|EPI_ISL_645681\|20 | ···················· | ···················· | ···················· | ···················· | ···················· | ···················· |
| R | 4534 hCoV-19/England/ALDP-B1BFBF/2020\|EPI_ISL_645682\|20 | ···················· | ···················· | ···················· | ···················· | ···················· | ···················· |
| R | 4535 hCoV-19/England/ALDP-B19FEE/2020\|EPI_ISL_645683\|20 | ···················· | ···················· | ···················· | ···················· | ······T············· | ···················· |
| R | 4536 hCoV-19/England/ALDP-B19F39/2020\|EPI_ISL_645684\|20 | ···················· | ···················· | ···················· | ···················· | ······T············· | ···················· |
| R | 4537 hCoV-19/England/ALDP-B1B517/2020\|EPI_ISL_645685\|20 | ···················· | ···················· | ···················· | ···················· | ···················· | ···················· |
| R | 4538 hCoV-19/England/ALDP-B1A0EA/2020\|EPI_ISL_645686\|20 | ···················· | ···················· | ···················· | ···················· | ···················· | ···················· |
| R | 4539 hCoV-19/England/ALDP-B1B429/2020\|EPI_ISL_645687\|20 | ···················· | ···················· | ···················· | ···················· | ···················· | ···················· |
| R | 4540 hCoV-19/England/ALDP-B1BB24/2020\|EPI_ISL_645688\|20 | ···················· | ···················· | ···················· | ···················· | ···················· | ···················· |
| R | 4541 hCoV-19/England/ALDP-B1CEFD/2020\|EPI_ISL_645690\|20 | ···················· | ···················· | ···················· | ···················· | ···················· | ···················· |
| R | 4542 hCoV-19/England/ALDP-B1AFDE/2020\|EPI_ISL_645691\|20 | ···················· | ···················· | ···················· | ···················· | ···················· | ···················· |
| R | 4543 hCoV-19/England/ALDP-B1A0CC/2020\|EPI_ISL_645692\|20 | ···················· | ···················· | ···················· | ···················· | ···················· | ···················· |
| R | 4544 hCoV-19/England/ALDP-B1B975/2020\|EPI_ISL_645693\|20 | ···················· | ···················· | ···················· | ···················· | ···················· | ···················· |
| R | 4545 hCoV-19/England/ALDP-B1BA90/2020\|EPI_ISL_645694\|20 | ···················· | ···················· | ···················· | ···················· | ···················· | ···················· |
| R | 4546 hCoV-19/England/ALDP-B1B070/2020\|EPI_ISL_645695\|20 | ···················· | ···················· | ···················· | ···················· | ···················· | ···················· |
| R | 4547 hCoV-19/England/ALDP-B19EF0/2020\|EPI_ISL_645696\|20 | ···················· | ···················· | ···················· | ···················· | ···················· | ···················· |
| R | 4548 hCoV-19/England/ALDP-B1BFCE/2020\|EPI_ISL_645697\|20 | ···················· | ···················· | ···················· | ···················· | ···················· | ···················· |
| R | 4549 hCoV-19/England/ALDP-B1BA63/2020\|EPI_ISL_645698\|20 | ···················· | ···················· | ···················· | ···················· | ···················· | ···A················ |
| R | 4550 hCoV-19/England/ALDP-B24627/2020\|EPI_ISL_646900\|20 | ···················· | ···················· | ···················· | ···················· | ···················· | ···················· |
| R | 4551 hCoV-19/England/ALDP-B24AFE/2020\|EPI_ISL_646901\|20 | ···················· | ···················· | ···················· | ···················· | ···················· | ···················· |
| R | 4552 hCoV-19/England/QEUH-B2DD56/2020\|EPI_ISL_646902\|20 | ···················· | ···················· | ···················· | ···················· | ···················· | ···················· |
| R | 4553 hCoV-19/England/ALDP-B24566/2020\|EPI_ISL_646903\|20 | ···················· | ···················· | ···················· | ···················· | ···················· | ···················· |
| R | 4554 hCoV-19/England/ALDP-B2392F/2020\|EPI_ISL_646904\|20 | ···················· | ···················· | ···················· | ···················· | ···················· | ···················· |
| R | 4555 hCoV-19/England/QEUH-B2DC68/2020\|EPI_ISL_646905\|20 | ···················· | ···················· | ···················· | ···················· | ···················· | ···A················ |
| R | 4556 hCoV-19/England/ALDP-B2445A/2020\|EPI_ISL_646906\|20 | ···················· | ···················· | ···················· | ···················· | ···················· | ···················· |
| R | 4557 hCoV-19/England/ALDP-B2487C/2020\|EPI_ISL_646907\|20 | ···················· | ···················· | ···················· | ···················· | ···················· | ···················· |
| R | 4558 hCoV-19/England/ALDP-B242BA/2020\|EPI_ISL_646908\|20 | ···················· | ···················· | ···················· | ···················· | ···················· | ···················· |
| R | 4559 hCoV-19/England/ALDP-B2387D/2020\|EPI_ISL_646909\|20 | ···················· | ···················· | ···················· | ···················· | ···················· | ···················· |
| R | 4560 hCoV-19/England/ALDP-B23B1A/2020\|EPI_ISL_646910\|20 | ···················· | ···················· | ···················· | ···················· | ···················· | ···················· |
| R | 4561 hCoV-19/England/ALDP-B2438A/2020\|EPI_ISL_646911\|20 | ···················· | ···················· | ···················· | ···················· | ···················· | ···················· |
| R | 4562 hCoV-19/England/QEUH-B2DAD7/2020\|EPI_ISL_646912\|20 | ···················· | ···················· | ···················· | ···················· | ···················· | ···················· |
| R | 4563 hCoV-19/England/ALDP-B23A1D/2020\|EPI_ISL_646913\|20 | ···················· | ···················· | ···················· | ···················· | ···················· | ···················· |
| R | 4564 hCoV-19/England/ALDP-B2397A/2020\|EPI_ISL_646914\|20 | ···················· | ···················· | ···················· | ···················· | ···················· | ···················· |
| R | 4565 hCoV-19/England/QEUH-B2D7D0/2020\|EPI_ISL_646915\|20 | ···················· | ···················· | ···················· | ···················· | ···················· | ···················· |
| R | 4566 hCoV-19/England/ALDP-B24539/2020\|EPI_ISL_646916\|20 | ···················· | ···················· | ···················· | ···················· | ···················· | ···················· |
| R | 4567 hCoV-19/England/ALDP-B24CDA/2020\|EPI_ISL_646917\|20 | ···················· | ···················· | ···················· | ···················· | ···················· | ···················· |
| R | 4568 hCoV-19/England/ALDP-B245DF/2020\|EPI_ISL_646918\|20 | ···················· | ···················· | ···················· | ···················· | ···················· | ···················· |
| R | 4569 hCoV-19/England/ALDP-B24399/2020\|EPI_ISL_646920\|20 | ···················· | ···················· | ···················· | ···················· | ···················· | ···················· |
| R | 4570 hCoV-19/England/QEUH-B2D7EF/2020\|EPI_ISL_646921\|20 | ···················· | ···················· | ···················· | ···················· | ···················· | ···················· |
| R | 4571 hCoV-19/England/ALDP-B24153/2020\|EPI_ISL_646922\|20 | ···················· | ···················· | ···················· | ···················· | ···················· | ···················· |
| R | 4572 hCoV-19/England/ALDP-B2484F/2020\|EPI_ISL_646924\|20 | ···················· | ···················· | ···················· | ···················· | ···················· | ···················· |
| R | 4573 hCoV-19/England/ALDP-B247BB/2020\|EPI_ISL_646925\|20 | ···················· | ···················· | ···················· | ···················· | ···················· | ···A················ |
| R | 4574 hCoV-19/England/ALDP-B24302/2020\|EPI_ISL_646926\|20 | ···················· | ···················· | ···················· | ···················· | ···················· | ···················· |
| R | 4575 hCoV-19/England/ALDP-B24830/2020\|EPI_ISL_646927\|20 | ···················· | ···················· | ···················· | ···················· | ···················· | ···················· |
| R | 4576 hCoV-19/England/ALDP-B23725/2020\|EPI_ISL_646928\|20 | ···················· | ···················· | ···················· | ···················· | ···················· | ···················· |
| R | 4577 hCoV-19/England/QEUH-B2D80A/2020\|EPI_ISL_646929\|20 | ···················· | ···················· | ···················· | ···················· | ···················· | ···················· |
| R | 4578 hCoV-19/England/ALDP-B247CA/2020\|EPI_ISL_646930\|20 | ···················· | ···················· | ···················· | ···················· | ···················· | ···················· |
| R | 4579 hCoV-19/England/ALDP-B249E2/2020\|EPI_ISL_646932\|20 | ···················· | ···················· | ···················· | ···················· | ···················· | ···················· |
| R | 4580 hCoV-19/England/ALDP-B242C9/2020\|EPI_ISL_646933\|20 | ···················· | ···················· | ···················· | ···················· | ···················· | ···················· |
| R | 4581 hCoV-19/England/ALDP-B249B5/2020\|EPI_ISL_646945\|20 | ···················· | ···················· | ···················· | ···················· | ···················· | ···················· |
| R | 4582 hCoV-19/England/QEUH-B2DAB9/2020\|EPI_ISL_646948\|20 | ···················· | ···················· | ···················· | ···················· | ···················· | ···················· |
| R | 4583 hCoV-19/England/ALDP-B243A8/2020\|EPI_ISL_646950\|20 | ···················· | ···················· | ···················· | ···················· | ···················· | ···················· |
| R | 4584 hCoV-19/England/ALDP-B2485E/2020\|EPI_ISL_646951\|20 | ···················· | ···················· | ···················· | ···················· | ···················· | ···················· |
| R | 4585 hCoV-19/England/ALDP-B245A2/2020\|EPI_ISL_646952\|20 | ···················· | ···················· | ···················· | ···················· | ···················· | ···················· |

| | | | | | | | |
|---|---|---|---|---|---|---|---|
| R | 4586 hCoV-19/England/ALDP-B2477F/2020\|EPI_ISL_646953\|20 | ···················· | ···················· | ···················· | ···················· | ···················· | ···················· |
| R | 4587 hCoV-19/England/ALDP-B23752/2020\|EPI_ISL_646954\|20 | ···················· | ···················· | ···················· | ···················· | ···················· | ···················· |
| R | 4588 hCoV-19/England/QEUH-B2DAAA/2020\|EPI_ISL_646955\|20 | ···················· | ···················· | ···················· | ···················· | ···················· | ···················· |
| R | 4589 hCoV-19/England/ALDP-B238F5/2020\|EPI_ISL_646978\|20 | ···················· | ···················· | ···················· | ···················· | ···················· | ···················· |
| R | 4590 hCoV-19/England/ALDP-B23B0B/2020\|EPI_ISL_646979\|20 | ···················· | ···················· | ···················· | ···················· | ···················· | ···················· |
| R | 4591 hCoV-19/England/ALDP-B2491F/2020\|EPI_ISL_646980\|20 | ···················· | ···················· | ···················· | ···················· | ···················· | ···················· |
| R | 4592 hCoV-19/England/ALDP-B24311/2020\|EPI_ISL_646982\|20 | ···················· | ···················· | ···················· | ···················· | ···················· | ···················· |
| R | 4593 hCoV-19/England/QEUH-B2DB6B/2020\|EPI_ISL_646983\|20 | ···················· | ···················· | ···················· | ···················· | ···················· | ···················· |
| R | 4594 hCoV-19/England/ALDP-B24171/2020\|EPI_ISL_646984\|20 | ···················· | ···················· | ···················· | ···················· | ···················· | ···················· |
| R | 4595 hCoV-19/England/ALDP-B23C44/2020\|EPI_ISL_646985\|20 | ···················· | ···················· | ···················· | ···················· | ···················· | ···················· |
| R | 4596 hCoV-19/England/ALDP-B238D7/2020\|EPI_ISL_646986\|20 | ···················· | ·········T·········· | ···················· | ···················· | ···················· | ···················· |
| R | 4597 hCoV-19/England/ALDP-B24760/2020\|EPI_ISL_646987\|20 | ···················· | ···················· | ···················· | ···················· | ···················· | ···················· |
| R | 4598 hCoV-19/England/QEUH-B2DA04/2020\|EPI_ISL_646988\|20 | ···················· | ···················· | ···················· | ···················· | ···················· | ···················· |
| R | 4599 hCoV-19/England/ALDP-B236BF/2020\|EPI_ISL_646990\|20 | ···················· | ···················· | ···················· | ···················· | ···················· | ···················· |
| R | 4600 hCoV-19/England/ALDP-B237CB/2020\|EPI_ISL_646991\|20 | ···················· | ···················· | ···················· | ···················· | ···················· | ··A················· |
| R | 4601 hCoV-19/England/QEUH-B2ECB2/2020\|EPI_ISL_646992\|20 | ···················· | ···················· | ···················· | ···················· | ···················· | ···················· |
| R | 4602 hCoV-19/England/ALDP-B24593/2020\|EPI_ISL_646993\|20 | ···················· | ···················· | ···················· | ···················· | ···················· | ···················· |
| R | 4603 hCoV-19/England/ALDP-B24821/2020\|EPI_ISL_646994\|20 | ···················· | ···················· | ···················· | ···················· | ···················· | ···················· |
| R | 4604 hCoV-19/England/ALDP-B237F8/2020\|EPI_ISL_646995\|20 | ···················· | ···················· | ···················· | ···················· | ···················· | ···················· |
| R | 4605 hCoV-19/England/QEUH-B2ECC1/2020\|EPI_ISL_646996\|20 | ···················· | ···················· | ···················· | ···················· | ···················· | ···················· |
| R | 4606 hCoV-19/England/ALDP-B236A0/2020\|EPI_ISL_646997\|20 | ···················· | ···················· | ···················· | ···················· | ···················· | ···················· |
| R | 4607 hCoV-19/England/ALDP-B24654/2020\|EPI_ISL_646998\|20 | ···················· | ·A·················· | ···················· | ···················· | ···················· | ··A················· |
| R | 4608 hCoV-19/England/ALDP-B241AE/2020\|EPI_ISL_646999\|20 | ···················· | ···················· | ···················· | ···················· | ···················· | ···················· |
| R | 4609 hCoV-19/Australia/QLD1148/2020\|EPI_ISL_639751\|2020 | ···················· | ···················· | ···················· | ···················· | ···················· | ···················· |
| R | 4610 hCoV-19/Australia/QLD1149/2020\|EPI_ISL_639752\|2020 | ···················· | ···················· | ···················· | ···················· | ···················· | ···················· |
| R | 4611 hCoV-19/Australia/QLD1150/2020\|EPI_ISL_639753\|2020 | ···················· | ···················· | ···················· | ···················· | ···················· | ···················· |
| R | 4612 hCoV-19/Australia/QLD1151/2020\|EPI_ISL_639754\|2020 | ···················· | ···················· | ···················· | ···················· | ···················· | ···················· |
| R | 4613 hCoV-19/Australia/QLD1152/2020\|EPI_ISL_639755\|2020 | ···················· | ···················· | ···················· | ···················· | ···················· | ···················· |
| R | 4614 hCoV-19/Australia/QLD1153/2020\|EPI_ISL_639756\|2020 | ···················· | ···················· | ···················· | ···················· | ···················· | ···················· |
| R | 4615 hCoV-19/Australia/QLD1154/2020\|EPI_ISL_639757\|2020 | ···················· | ···················· | ···················· | ···················· | ···················· | ···················· |
| R | 4616 hCoV-19/Australia/QLD1155/2020\|EPI_ISL_639758\|2020 | ···················· | ···················· | ···················· | ···················· | ···················· | ···················· |
| R | 4617 hCoV-19/Australia/QLD1156/2020\|EPI_ISL_639759\|2020 | ···················· | ···················· | ···················· | ···················· | ···················· | ···················· |
| R | 4618 hCoV-19/Australia/QLD1157/2020\|EPI_ISL_639760\|2020 | ···················· | ···················· | ···················· | ···················· | ···················· | ···················· |
| R | 4619 hCoV-19/Australia/QLD1158/2020\|EPI_ISL_639761\|2020 | ···················· | ···················· | ···················· | ···················· | ···················· | ···················· |
| R | 4620 hCoV-19/Australia/QLD1160/2020\|EPI_ISL_639762\|2020 | ···················· | ···················· | ···················· | ···················· | ···················· | ···················· |
| R | 4621 hCoV-19/Australia/QLD1172/2020\|EPI_ISL_639763\|2020 | ···················· | ···················· | ···················· | ···················· | ···················· | ···················· |
| R | 4622 hCoV-19/Australia/QLD1173/2020\|EPI_ISL_639764\|2020 | ···················· | ···················· | ···················· | ···················· | ···················· | ···················· |
| R | 4623 hCoV-19/Australia/QLD1174/2020\|EPI_ISL_639765\|2020 | ···················· | ···················· | ···················· | ···················· | ···················· | ···················· |
| R | 4624 hCoV-19/Australia/QLD1175/2020\|EPI_ISL_639766\|2020 | ···················· | ···················· | ···················· | ···················· | ···················· | ···················· |
| R | 4625 hCoV-19/Australia/QLD1176/2020\|EPI_ISL_639767\|2020 | ···················· | ···················· | ···················· | ···················· | ···················· | ···················· |
| R | 4626 hCoV-19/Australia/QLD1177/2020\|EPI_ISL_639768\|2020 | ···················· | ···················· | ···················· | ···················· | ···················· | ···················· |
| R | 4627 hCoV-19/Australia/QLD1178/2020\|EPI_ISL_639769\|2020 | ···················· | ···················· | ···················· | ···················· | ···················· | ···················· |
| R | 4628 hCoV-19/Australia/QLD1179/2020\|EPI_ISL_639770\|2020 | ···················· | ···················· | ···················· | ···················· | ···················· | ···················· |
| R | 4629 hCoV-19/Australia/QLD1180/2020\|EPI_ISL_639771\|2020 | ···················· | ···················· | ···················· | ···················· | ···················· | ···················· |
| R | 4630 hCoV-19/Australia/QLD1181/2020\|EPI_ISL_639772\|2020 | ···················· | ···················· | ···················· | ···················· | ···················· | ···················· |
| R | 4631 hCoV-19/Australia/QLD1182/2020\|EPI_ISL_639773\|2020 | ···················· | ·············A······ | ···················· | ···················· | ···················· | ···················· |
| R | 4632 hCoV-19/Australia/QLD1183/2020\|EPI_ISL_639774\|2020 | ···················· | ···················· | ···················· | ···················· | ···················· | ···················· |
| R | 4633 hCoV-19/Australia/QLD1184/2020\|EPI_ISL_639775\|2020 | ···················· | ···················· | ···················· | ···················· | ···················· | ···················· |
| R | 4634 hCoV-19/Australia/QLD1185/2020\|EPI_ISL_639776\|2020 | ···················· | ···················· | ···················· | ···················· | ···················· | ···················· |
| R | 4635 hCoV-19/Australia/QLD1186/2020\|EPI_ISL_639777\|2020 | ···················· | ···················· | ···················· | ···················· | ···················· | ···················· |
| R | 4636 hCoV-19/Australia/QLD1187/2020\|EPI_ISL_639778\|2020 | ···················· | ···················· | ···················· | ···················· | ···················· | ···················· |
| R | 4637 hCoV-19/Australia/QLD1188/2020\|EPI_ISL_639779\|2020 | ···················· | ···················· | ···················· | ···················· | ···················· | ···················· |
| R | 4638 hCoV-19/Australia/QLD1189/2020\|EPI_ISL_639780\|2020 | ···················· | ···················· | ···················· | ···················· | ···················· | ···················· |
| R | 4639 hCoV-19/Australia/QLD1190/2020\|EPI_ISL_639781\|2020 | ···················· | ···················· | ···················· | ···················· | ···················· | ···················· |
| R | 4640 hCoV-19/Australia/QLD1191/2020\|EPI_ISL_639782\|2020 | ···················· | ···················· | ···················· | ···················· | ···················· | ···················· |
| R | 4641 hCoV-19/Australia/QLD1192/2020\|EPI_ISL_639783\|2020 | ···················· | ···················· | ···················· | ···················· | ···················· | ···················· |
| R | 4642 hCoV-19/Australia/QLD1193/2020\|EPI_ISL_639784\|2020 | ···················· | ···················· | ···················· | ···················· | ···················· | ···················· |
| R | 4643 hCoV-19/Australia/QLD1194/2020\|EPI_ISL_639785\|2020 | ···················· | ···················· | ···················· | ···················· | ···················· | ···················· |
| R | 4644 hCoV-19/Australia/QLD1195/2020\|EPI_ISL_639786\|2020 | ···················· | ···················· | ···················· | ···················· | ···················· | ···················· |
| R | 4645 hCoV-19/Australia/QLD1196/2020\|EPI_ISL_639787\|2020 | ···················· | ···················· | ···················· | ···················· | ···················· | ···················· |
| R | 4646 hCoV-19/Australia/QLD1197/2020\|EPI_ISL_639788\|2020 | ···················· | ···················· | ···················· | ···················· | ···················· | ···················· |
| R | 4647 hCoV-19/Australia/QLD1198/2020\|EPI_ISL_639789\|2020 | ···················· | ···················· | ···················· | ···················· | ···················· | ···················· |
| R | 4648 hCoV-19/Australia/QLD1199/2020\|EPI_ISL_639790\|2020 | ···················· | ···················· | ···················· | ···················· | ···················· | ···················· |
| R | 4649 hCoV-19/Australia/QLD1200/2020\|EPI_ISL_639791\|2020 | ···················· | ···················· | ···················· | ···················· | ···················· | ···················· |
| R | 4650 hCoV-19/Australia/QLD1201/2020\|EPI_ISL_639792\|2020 | ···················· | ···················· | ···················· | ···················· | ···················· | ···················· |
| R | 4651 hCoV-19/Australia/QLD1202/2020\|EPI_ISL_639793\|2020 | ···················· | ···················· | ···················· | ···················· | ···················· | ···················· |
| R | 4652 hCoV-19/Australia/QLD1203/2020\|EPI_ISL_639794\|2020 | ···················· | ···················· | ···················· | ···················· | ···················· | ···················· |
| R | 4653 hCoV-19/Australia/QLD1204/2020\|EPI_ISL_639795\|2020 | ···················· | ···················· | ···················· | ···················· | ···················· | ···················· |
| R | 4654 hCoV-19/Australia/QLD1205/2020\|EPI_ISL_639796\|2020 | ···················· | ···················· | ···················· | ···················· | ···················· | ···················· |
| R | 4655 hCoV-19/Australia/QLD1206/2020\|EPI_ISL_639797\|2020 | ···················· | ···················· | ···················· | ···················· | ···················· | ···················· |
| R | 4656 hCoV-19/Australia/QLD1207/2020\|EPI_ISL_639798\|2020 | ···················· | ···················· | ···················· | ···················· | ···················· | ···················· |
| R | 4657 hCoV-19/Australia/QLD1208/2020\|EPI_ISL_639799\|2020 | ···················· | ···················· | ···················· | ···················· | ···················· | ···················· |
| R | 4658 hCoV-19/Australia/VIC16059/2020\|EPI_ISL_640703\|202 | ···················· | ···················· | ···················· | ···················· | ···················· | ···················· |
| R | 4660 hCoV-19/Australia/VIC15993/2020\|EPI_ISL_640706\|202 | ···················· | ···················· | ···················· | ···················· | ···················· | ···················· |
| R | 4661 hCoV-19/Australia/VIC16521/2020\|EPI_ISL_640707\|202 | ···················· | ···················· | ···················· | ···················· | ···················· | ···················· |
| R | 4662 hCoV-19/Australia/VIC16555/2020\|EPI_ISL_640708\|202 | ···················· | ···················· | ···················· | ···················· | ···················· | ···················· |
| R | 4663 hCoV-19/Australia/VIC16056/2020\|EPI_ISL_640709\|202 | ···················· | ···················· | ···················· | ···················· | ···················· | ···················· |
| R | 4664 hCoV-19/Australia/VIC12475/2020\|EPI_ISL_640710\|202 | ···················· | ···················· | ···················· | ···················· | ···················· | ···················· |
| R | 4665 hCoV-19/Australia/VIC16461/2020\|EPI_ISL_640711\|202 | ···················· | ···················· | ···················· | ···················· | ···················· | ···················· |
| R | 4666 hCoV-19/Australia/VIC13251/2020\|EPI_ISL_640713\|202 | ···················· | ···················· | ···················· | ···················· | ···················· | ···················· |

```
R 4667 hCoV-19/Australia/VIC12090/2020|EPI_ISL_640714|202 .................... .................... ..................... .................... .................... ....................
R 4668 hCoV-19/Australia/VIC16472/2020|EPI_ISL_640715|202 .................... .................... ..................... .................... .................... ....................
R 4669 hCoV-19/Australia/VIC12373/2020|EPI_ISL_640717|202 .................... .................... ..................... .................... .................... ....................
R 4670 hCoV-19/Australia/VIC16077/2020|EPI_ISL_640718|202 .................... .................... ..................... .................... .................... ....................
R 4671 hCoV-19/Australia/VIC16188/2020|EPI_ISL_640719|202 .................... .................... ..................... .................... .................... ....................
R 4673 hCoV-19/Australia/VIC12357/2020|EPI_ISL_640723|202 .................... .................... ..................... .................... .................... ....................
R 4674 hCoV-19/Australia/VIC12769/2020|EPI_ISL_640724|202 .................... .................... ..................... .................... .................... ....................
R 4675 hCoV-19/Australia/VIC11689/2020|EPI_ISL_640725|202 .................... .................... ..................... .................... .................... ....................
R 4676 hCoV-19/Australia/VIC12016/2020|EPI_ISL_640726|202 .................... .................... ..................... .................... .................... ....................
R 4677 hCoV-19/Australia/VIC16611/2020|EPI_ISL_640727|202 .................... .................... ..................... .................... .................... ..............N.....
R 4678 hCoV-19/Australia/VIC12638/2020|EPI_ISL_640728|202 .................... .................... ..................... .................... .................... ....................
R 4679 hCoV-19/Australia/VIC12287/2020|EPI_ISL_640729|202 .................... .................... ..................... .................... .................... ....................
R 4682 hCoV-19/Australia/VIC15994/2020|EPI_ISL_640756|202 .................... .................... ..................... .................... .................... ....................
R 4683 hCoV-19/Australia/VIC16126/2020|EPI_ISL_640757|202 .................... .................... ..................... .................... .................... ....................
R 4684 hCoV-19/Australia/VIC16531/2020|EPI_ISL_640758|202 .................... .................... ..................... .................... .................... ....................
R 4685 hCoV-19/Australia/VIC12856/2020|EPI_ISL_640759|202 .................... .................... ..................... .................... .................... ....................
R 4686 hCoV-19/Australia/VIC16481/2020|EPI_ISL_640760|202 .................... .................... ..................... .................... NNN................. .................... 1
R 4687 hCoV-19/Australia/VIC12854/2020|EPI_ISL_640761|202 .................... .................... ..................... .................... .................... ....................
R 4688 hCoV-19/Australia/VIC12630/2020|EPI_ISL_640762|202 .................... .................... ..................... .................... .................... ....................
R 4689 hCoV-19/Australia/VIC12377/2020|EPI_ISL_640765|202 .................... .................... ..................... .................... .................... ....................
R 4690 hCoV-19/Australia/VIC13242/2020|EPI_ISL_640766|202 .................... .................... ..................... .................... .................... ....................
R 4691 hCoV-19/Australia/VIC12422/2020|EPI_ISL_640767|202 .................... .................... ..................... .................... .................... ....................
R 4692 hCoV-19/Australia/VIC15991/2020|EPI_ISL_640770|202 .................... .................... ..................... .................... .................... ....................
R 4693 hCoV-19/Australia/VIC12820/2020|EPI_ISL_640773|202 .................... .................... ..................... .................... .................... ....................
R 4695 hCoV-19/Australia/VIC16338/2020|EPI_ISL_640777|202 .................... .................... ..................... .................... .................... ....................
R 4696 hCoV-19/Australia/VIC15918/2020|EPI_ISL_640778|202 .................... .................... ..................... .................... .................... ....................
R 4699 hCoV-19/Australia/VIC15936/2020|EPI_ISL_640782|202 .................... .................... ..................... .................... .................... ....................
R 4700 hCoV-19/Australia/VIC15870/2020|EPI_ISL_640784|202 .................... .................... ..................... .................... .................... ....................
R 4701 hCoV-19/Australia/VIC12570/2020|EPI_ISL_640785|202 .................... .................... ..................... .................... .................... ....................
R 4702 hCoV-19/Australia/VIC16075/2020|EPI_ISL_640786|202 .................... .................... ..................... .................... .................... ....................
R 4703 hCoV-19/Australia/VIC16507/2020|EPI_ISL_640788|202 .................... .................... ..................... .................... .................... ....................
R 4704 hCoV-19/Australia/VIC16663/2020|EPI_ISL_640789|202 .................... .................... ..................... .................... .................... ....................
R 4705 hCoV-19/Australia/VIC16091/2020|EPI_ISL_640790|202 .................... .................... ..................... .................... .................... ....................
R 4706 hCoV-19/Australia/VIC16071/2020|EPI_ISL_640792|202 .................... .................... ..................... .................... .................... ....................
R 4707 hCoV-19/Australia/VIC16547/2020|EPI_ISL_640793|202 .................... .................... ..................... .................... .................... ....................
R 4708 hCoV-19/Australia/VIC16637/2020|EPI_ISL_640794|202 .................... .................... ..................... .................... .................... ....................
R 4709 hCoV-19/Australia/VIC12917/2020|EPI_ISL_640796|202 .................... .................... ..................... .................... .................... ....................
R 4710 hCoV-19/Australia/VIC16523/2020|EPI_ISL_640798|202 .................... .................... ..................... .................... .................... ....................
R 4711 hCoV-19/Australia/VIC16130/2020|EPI_ISL_640799|202 .................... .................... ..................... .................... .................... ....................
R 4712 hCoV-19/Ireland/WW-NVRL-AIIDV0277v1/2020|EPI_ISL_6 .................... .................... ..................... .................... .................... ....................
R 4713 hCoV-19/Ireland/D-NVRL-AIIDV0216v1/2020|EPI_ISL_64 .................... .................... ..................... .................... .................... ....................
R 4714 hCoV-19/Ireland/WW-NVRL-AIIDV0082v1/2020|EPI_ISL_6 .................... .................... ..................... .................... .................... ....................
R 4715 hCoV-19/Ireland/WW-NVRL-AIIDV0082v2/2020|EPI_ISL_6 .................... .................... ..................... .................... .................... ....................
R 4716 hCoV-19/Ireland/D-NVRL-AIIDV0328v1/2020|EPI_ISL_64 .................... .................... ..................... .................... .................... ....................
R 4717 hCoV-19/Ireland/D-NVRL-AIIDV0040v1/2020|EPI_ISL_64 .................... .................... ..................... .................... .................... ....................
R 4718 hCoV-19/Ireland/D-NVRL-AIIDV0081v2/2020|EPI_ISL_64 .................... .................... ..................... .................... .................... ....................
R 4719 hCoV-19/Ireland/D-NVRL-AIIDV0432v4/2020|EPI_ISL_64 .................... .................... ..................... .................... .................... ....................
R 4720 hCoV-19/Ireland/D-NVRL-AIIDV0141v1/2020|EPI_ISL_64 .................... ...................A ..................... .................... .................... .................... 1
R 4721 hCoV-19/Ireland/D-NVRL-AIIDV0141v2/2020|EPI_ISL_64 .................... ...................A ..................... .................... .................... .................... 1
R 4722 hCoV-19/Ireland/D-NVRL-AIIDV0102v1/2020|EPI_ISL_64 .................... .................A.. ..................... .................... .................... .................... 1
R 4723 hCoV-19/Ireland/WW-NVRL-AIIDV0099v1/2020|EPI_ISL_6 .................... .................... ..................... .................... .................... ....................
R 4724 hCoV-19/Ireland/D-NVRL-AIIDV0432v5/2020|EPI_ISL_64 .................... .................... ..................... .................... .................... ....................
R 4725 hCoV-19/Ireland/D-NVRL-AIIDV0192v2/2020|EPI_ISL_64 .................... .................... ..................... .................... .................... ....................
R 4726 hCoV-19/Ireland/D-NVRL-AIIDV0264v1/2020|EPI_ISL_64 .................... .................... ..................... .................... .................... ....................
R 4727 hCoV-19/Ireland/WW-NVRL-AIIDV0258v1/2020|EPI_ISL_6 .................... .................... ..................... .................... .................... ....................
R 4728 hCoV-19/Ireland/D-NVRL-AIIDV0072v3/2020|EPI_ISL_64 .................... .................... ..................... .................... .................... ....................
R 4729 hCoV-19/Ireland/D-NVRL-AIIDV0264v2/2020|EPI_ISL_64 .................... .................... ..................... .................... .................... ....................
R 4730 hCoV-19/Ireland/WW-NVRL-AIIDV0486v1/2020|EPI_ISL_6 .................... .................... ..................... .................... .................... ....................
R 4731 hCoV-19/Ireland/WW-NVRL-AIIDV0277v2/2020|EPI_ISL_6 .................... .................... ..................... .................... .................... ....................
R 4732 hCoV-19/England/CAMC-B3757F/2020|EPI_ISL_645500|20 .................... .................... ..................... .................... .................... ....................
R 4733 hCoV-19/England/ALDP-B3EA4C/2020|EPI_ISL_645502|20 .................... .................... ..................... .................... .................... ....................
R 4734 hCoV-19/England/ALDP-B3E903/2020|EPI_ISL_645503|20 .................... .................... ..................... .................... .................... ....................
R 4735 hCoV-19/England/CAMC-B36631/2020|EPI_ISL_645505|20 .................... .................... ..................... .................... .................... ....................
R 4736 hCoV-19/England/CAMC-B37427/2020|EPI_ISL_645506|20 .................... .................... ..................... .................... .................... ....................
R 4737 hCoV-19/England/CAMC-B37269/2020|EPI_ISL_645507|20 .................... .................... ..................... .................... ......T............. ....................
R 4738 hCoV-19/England/ALDP-B3E51E/2020|EPI_ISL_645508|20 .................... .................... ..................... .................... .................... ....................
R 4739 hCoV-19/England/CAMC-B3617C/2020|EPI_ISL_645509|20 .................... .................... ..................... .................... .................... ....................
R 4740 hCoV-19/England/CAMC-B35E67/2020|EPI_ISL_645510|20 .................... .A.................. ..................... .................... .................... ...A................
R 4741 hCoV-19/England/CAMC-B373EE/2020|EPI_ISL_645511|20 .................... .................... ..................... .................... .................... ....................
R 4742 hCoV-19/England/CAMC-B3677A/2020|EPI_ISL_645512|20 .................... .................... ..................... .................... .................... ....................
R 4743 hCoV-19/England/MILK-B3D227/2020|EPI_ISL_645513|20 .................... .................... ..................... .................... .................... ....................
R 4744 hCoV-19/England/CAMC-B3608E/2020|EPI_ISL_645514|20 .................... .................... ..................... .................... .................... ....................
R 4745 hCoV-19/England/CAMC-B360D9/2020|EPI_ISL_645515|20 .................... .................... ..................... .................... .................... ....................
R 4746 hCoV-19/England/CAMC-B36024/2020|EPI_ISL_645516|20 .................... .................... ..................... .................... .................... ....................
R 4747 hCoV-19/England/MILK-B3E4A8/2020|EPI_ISL_645517|20 .................... .A.................. ..................... .................... .................... ...A................
R 4748 hCoV-19/England/CAMC-B3631C/2020|EPI_ISL_645519|20 .................... .................... ..................... .................... .................... ....................
R 4749 hCoV-19/England/CAMC-B37621/2020|EPI_ISL_645521|20 .................... .................... ..................... .................... .................... ....................
R 4750 hCoV-19/England/CAMC-B362A6/2020|EPI_ISL_645522|20 .................... .................... ..................... .................... .................... ....................
R 4751 hCoV-19/England/CAMC-B36394/2020|EPI_ISL_645523|20 .................... .................... ..................... .................... .................... ....................
R 4752 hCoV-19/England/CAMC-B36947/2020|EPI_ISL_645524|20 .................... .................... ..................... .................... .................... ....................
```

| | | | | | | | |
|---|---|---|---|---|---|---|---|
| R | 4753 hCoV-19/England/CAMC-B367A7/2020\|EPI_ISL_645525\|20 | ···················· | ···················· | ····················· | ···················· | ···················· | ···················· |
| R | 4754 hCoV-19/England/ALDP-B24609/2020\|EPI_ISL_646800\|20 | ···················· | ···················· | ····················· | ···················· | ···················· | ···················· |
| R | 4755 hCoV-19/England/ALDP-B248C7/2020\|EPI_ISL_646801\|20 | ···················· | ···················· | ····················· | ···················· | ···················· | ···················· |
| R | 4756 hCoV-19/England/ALDP-B23831/2020\|EPI_ISL_646802\|20 | ···················· | ···················· | ····················· | ···················· | ···················· | ···················· |
| R | 4757 hCoV-19/England/ALDP-B23A4A/2020\|EPI_ISL_646803\|20 | ···················· | ···················· | ····················· | ···················· | ···················· | ···················· |
| R | 4758 hCoV-19/England/ALDP-B243F3/2020\|EPI_ISL_646804\|20 | ···················· | ···················· | ····················· | ···················· | ···················· | ···················· |
| R | 4759 hCoV-19/England/ALDP-B237BC/2020\|EPI_ISL_646805\|20 | ···················· | ···················· | ····················· | ···················· | ···················· | ···················· |
| R | 4760 hCoV-19/England/ALDP-B243C6/2020\|EPI_ISL_646806\|20 | ···················· | ···················· | ····················· | ···················· | ···················· | ···················· |
| R | 4761 hCoV-19/England/ALDP-B2442D/2020\|EPI_ISL_646807\|20 | ···················· | ···················· | ····················· | ···················· | ···················· | ···················· |
| R | 4762 hCoV-19/England/ALDP-B23901/2020\|EPI_ISL_646808\|20 | ···················· | ···················· | ····················· | ···················· | ···················· | ···················· |
| R | 4763 hCoV-19/England/ALDP-B23707/2020\|EPI_ISL_646811\|20 | ···················· | ···················· | ····················· | ···················· | ···················· | ···················· |
| R | 4764 hCoV-19/England/ALDP-B23910/2020\|EPI_ISL_646812\|20 | ···················· | ···················· | ····················· | ···················· | ···················· | ···················· |
| R | 4765 hCoV-19/England/ALDP-B241F9/2020\|EPI_ISL_646813\|20 | ···················· | ···················· | ····················· | ···················· | ···················· | ···················· |
| R | 4766 hCoV-19/England/ALDP-B23AC2/2020\|EPI_ISL_646814\|20 | ···················· | ···················· | ····················· | ···················· | ···················· | ···A················ |
| R | 4767 hCoV-19/England/ALDP-B24B19/2020\|EPI_ISL_646815\|20 | ···················· | ···················· | ····················· | ···················· | ···················· | ···················· |
| R | 4768 hCoV-19/England/ALDP-B241DB/2020\|EPI_ISL_646817\|20 | ···················· | ···················· | ····················· | ···················· | ···················· | ···················· |
| R | 4769 hCoV-19/England/QEUH-B2DCD1/2020\|EPI_ISL_646818\|20 | ···················· | ···················· | ····················· | ···················· | ···················· | ···················· |
| R | 4770 hCoV-19/England/ALDP-B246CD/2020\|EPI_ISL_646819\|20 | ···················· | ···················· | ····················· | ···················· | ···················· | ···················· |
| R | 4771 hCoV-19/England/ALDP-B24742/2020\|EPI_ISL_646820\|20 | ···················· | ···················· | ····················· | ···················· | ···················· | ···················· |
| R | 4772 hCoV-19/England/ALDP-B24A67/2020\|EPI_ISL_646822\|20 | ···················· | ···················· | ····················· | ···················· | ···················· | ···················· |
| R | 4773 hCoV-19/England/ALDP-B24C43/2020\|EPI_ISL_646823\|20 | ···················· | ···················· | ····················· | ···················· | ···················· | ···················· |
| R | 4774 hCoV-19/England/ALDP-B2395C/2020\|EPI_ISL_646824\|20 | ···················· | ···················· | ····················· | ···················· | ···················· | ···················· |
| R | 4775 hCoV-19/England/QEUH-B2DAE6/2020\|EPI_ISL_646825\|20 | ···················· | ···················· | ····················· | ···················· | ···················· | ···················· |
| R | 4776 hCoV-19/England/QEUH-B2DB98/2020\|EPI_ISL_646826\|20 | ···················· | ···················· | ····················· | ···················· | ···················· | ···················· |
| R | 4777 hCoV-19/England/ALDP-B24A85/2020\|EPI_ISL_646827\|20 | ···················· | ···················· | ····················· | ···················· | ···················· | ···················· |
| R | 4778 hCoV-19/England/ALDP-B24A1C/2020\|EPI_ISL_646828\|20 | ···················· | ·········T·········· | ····················· | ···················· | ···················· | ···················· |
| R | 4779 hCoV-19/England/ALDP-B246FA/2020\|EPI_ISL_646829\|20 | ···················· | ···················· | ····················· | ···················· | ···················· | ···················· |
| R | 4780 hCoV-19/England/ALDP-B24223/2020\|EPI_ISL_646830\|20 | ···················· | ···················· | ····················· | ···················· | ···················· | ···················· |
| R | 4781 hCoV-19/England/ALDP-B24D04/2020\|EPI_ISL_646831\|20 | ···················· | ···················· | ····················· | ···················· | ···················· | ···················· |
| R | 4782 hCoV-19/England/ALDP-B273F0/2020\|EPI_ISL_646832\|20 | ···················· | ·A·················· | ····················· | ···················· | ···················· | ···A················ |
| R | 4783 hCoV-19/England/ALDP-B242AB/2020\|EPI_ISL_646833\|20 | ···················· | ···················· | ····················· | ···················· | ···················· | ···················· |
| R | 4784 hCoV-19/England/ALDP-B244F0/2020\|EPI_ISL_646834\|20 | ···················· | ···················· | ····················· | ···················· | ···················· | ···················· |
| R | 4785 hCoV-19/England/ALDP-B2493D/2020\|EPI_ISL_646835\|20 | ···················· | ···················· | ····················· | ···················· | ···················· | ···················· |
| R | 4786 hCoV-19/England/ALDP-B2496A/2020\|EPI_ISL_646836\|20 | ···················· | ···················· | ····················· | ···················· | ···················· | ···················· |
| R | 4787 hCoV-19/England/QEUH-B2ECFE/2020\|EPI_ISL_646837\|20 | ···················· | ···················· | ····················· | ···················· | ···················· | ···················· |
| R | 4788 hCoV-19/England/ALDP-B24672/2020\|EPI_ISL_646838\|20 | ···················· | ···················· | ····················· | ···················· | ···················· | ···················· |
| R | 4789 hCoV-19/England/ALDP-B236CE/2020\|EPI_ISL_646839\|20 | ···················· | ···················· | ····················· | ···················· | ···················· | ···················· |
| R | 4790 hCoV-19/England/ALDP-B23A95/2020\|EPI_ISL_646841\|20 | ···················· | ···················· | ····················· | ···················· | ···················· | ···················· |
| R | 4791 hCoV-19/England/ALDP-B24CF8/2020\|EPI_ISL_646842\|20 | ···················· | ·A·················· | ····················· | ···················· | ···················· | ···A················ |
| R | 4792 hCoV-19/England/ALDP-B2488B/2020\|EPI_ISL_646843\|20 | ···················· | ···················· | ····················· | ···················· | ···················· | ···················· |
| R | 4793 hCoV-19/England/ALDP-B24690/2020\|EPI_ISL_646844\|20 | ···················· | ···················· | ····················· | ···················· | ···················· | ···················· |
| R | 4794 hCoV-19/England/ALDP-B2434E/2020\|EPI_ISL_646845\|20 | ···················· | ···················· | ····················· | ···················· | ···················· | ···················· |
| R | 4795 hCoV-19/England/ALDP-B24A76/2020\|EPI_ISL_646846\|20 | ···················· | ···················· | ····················· | ···················· | ···················· | ···················· |
| R | 4796 hCoV-19/England/ALDP-B24900/2020\|EPI_ISL_646847\|20 | ···················· | ···················· | ····················· | ···················· | ···················· | ···················· |
| R | 4797 hCoV-19/Wales/ALDP-B24C9E/2020\|EPI_ISL_646848\|2020 | ···················· | ···················· | ····················· | ···················· | ···················· | ···················· |
| R | 4798 hCoV-19/England/ALDP-B2437B/2020\|EPI_ISL_646849\|20 | ···················· | ···················· | ····················· | ···················· | ···················· | ···················· |
| R | 4799 hCoV-19/England/QEUH-B2D7C1/2020\|EPI_ISL_646850\|20 | ···················· | ···················· | ····················· | ···················· | ···················· | ···················· |
| R | 4800 hCoV-19/England/ALDP-B24232/2020\|EPI_ISL_646851\|20 | ···················· | ···················· | ····················· | ···················· | ···················· | ···················· |
| R | 4801 hCoV-19/England/ALDP-B247F7/2020\|EPI_ISL_646852\|20 | ···················· | ···················· | ····················· | ···················· | ···················· | ···A················ |
| R | 4802 hCoV-19/England/QEUH-B2DAC8/2020\|EPI_ISL_646853\|20 | ···················· | ···················· | ····················· | ···················· | ···················· | ···················· |
| R | 4803 hCoV-19/England/QEUH-B2DC1D/2020\|EPI_ISL_646854\|20 | ···················· | ·A·················· | ····················· | ···················· | ···················· | ···················· |
| R | 4804 hCoV-19/England/ALDP-B23664/2020\|EPI_ISL_646855\|20 | ···················· | ···················· | ····················· | ···················· | ···················· | ···················· |
| R | 4805 hCoV-19/England/ALDP-B237E9/2020\|EPI_ISL_646856\|20 | ···················· | ···················· | ····················· | ···················· | ···················· | ···················· |
| R | 4806 hCoV-19/USA/CA-ALSR-4473/2020\|EPI_ISL_649050\|2020- | ···················· | ···················· | ····················· | ···················· | ···················· | ···················· |
| R | 4807 hCoV-19/USA/CA-ALSR-4474/2020\|EPI_ISL_649051\|2020- | ···················· | ·A·················· | ····················· | ···················· | ···················· | ···················· |
| R | 4808 hCoV-19/USA/CA-ALSR-4475/2020\|EPI_ISL_649052\|2020- | ···················· | ···················· | ····················· | ···················· | ···················· | ···················· |
| R | 4809 hCoV-19/USA/CA-ALSR-4478/2020\|EPI_ISL_649053\|2020- | ···················· | ·A·················· | ····················· | ···················· | ···················· | ···················· |
| R | 4810 hCoV-19/USA/CA-ALSR-4479/2020\|EPI_ISL_649054\|2020- | ···················· | ·A·················· | ····················· | ···················· | ···················· | ···················· |
| R | 4811 hCoV-19/USA/CA-ALSR-4481/2020\|EPI_ISL_649055\|2020- | ···················· | ·A·················· | ····················· | ···················· | ···················· | ···················· |
| R | 4812 hCoV-19/USA/CA-ALSR-4484/2020\|EPI_ISL_649056\|2020- | ···················· | ·A·················· | ····················· | ···················· | ···················· | ···················· |
| R | 4816 hCoV-19/Israel/CVL-n2899/2020\|EPI_ISL_649066\|2020- | ···················· | ···················· | ····················· | ···················· | ···················· | ···················· |
| R | 4817 hCoV-19/Israel/CVL-n1962/2020\|EPI_ISL_649067\|2020- | ···················· | ···················· | ····················· | ···················· | ···················· | ···················· |
| R | 4818 hCoV-19/Israel/CVL-n3837/2021\|EPI_ISL_649068\|2020- | ···················· | ···················· | ····················· | ···················· | ···················· | ···················· |
| R | 4819 hCoV-19/Israel/CVL-n2125/2020\|EPI_ISL_649069\|2020- | ···················· | ···················· | ····················· | ···················· | ···················· | ···················· |
| R | 4820 hCoV-19/Israel/CVL-n3953/2022\|EPI_ISL_649070\|2020- | ···················· | ···················· | ····················· | ···················· | ···················· | ···················· |
| R | 4821 hCoV-19/Israel/CVL-n4097/2029\|EPI_ISL_649071\|2020- | ···················· | ···················· | ····················· | ···················· | ···················· | ···················· |
| R | 4822 hCoV-19/Israel/CVL-n4109/2030\|EPI_ISL_649072\|2020- | ···················· | ···················· | ····················· | ···················· | ···················· | ···················· |
| R | 4823 hCoV-19/Israel/CVL-n2820/2020\|EPI_ISL_649073\|2020- | ···················· | ···················· | ····················· | ···················· | ···················· | ···················· |
| R | 4824 hCoV-19/Israel/CVL-n3021/2020\|EPI_ISL_649074\|2020- | ···················· | ···················· | ····················· | ···················· | ···················· | ···················· |
| R | 4825 hCoV-19/Israel/CVL-n3380/2020\|EPI_ISL_649075\|2020- | ···················· | ···················· | ····················· | ···················· | ···················· | ···················· |
| R | 4826 hCoV-19/Israel/CVL-n3020/2020\|EPI_ISL_649076\|2020- | ···················· | ···················· | ····················· | ···················· | ···················· | ···················· |
| R | 4827 hCoV-19/Israel/CVL-n3607/2020\|EPI_ISL_649077\|2020- | ···················· | ···················· | ····················· | ···················· | ···················· | ···················· |
| R | 4828 hCoV-19/Israel/CVL-n4014/2024\|EPI_ISL_649078\|2020- | ···················· | ···················· | ····················· | ···················· | ···················· | ···················· |
| R | 4829 hCoV-19/Israel/CVL-n3304/2020\|EPI_ISL_649079\|2020- | ···················· | ···················· | ····················· | ···················· | ···················· | ···················· |
| R | 4830 hCoV-19/Israel/CVL-n2888/2020\|EPI_ISL_649080\|2020- | ···················· | ···················· | ····················· | ···················· | ···················· | ···················· |
| R | 4831 hCoV-19/Israel/CVL-n3862/2037\|EPI_ISL_649081\|2020- | ···················· | ···················· | ····················· | ···················· | ···················· | ···················· |
| R | 4832 hCoV-19/Israel/CVL-n3767/2020\|EPI_ISL_649082\|2020- | ···················· | ···················· | ····················· | ···················· | ···················· | ···················· |
| R | 4833 hCoV-19/Israel/CVL-n2875/2020\|EPI_ISL_649083\|2020- | ···················· | ···················· | ····················· | ···················· | ···················· | ···················· |
| R | 4835 hCoV-19/Israel/CVL-n4414/2035\|EPI_ISL_649085\|2020- | ···················· | ···················· | ····················· | ···················· | ···················· | ···················· |
| R | 4836 hCoV-19/Israel/CVL-n2494/2020\|EPI_ISL_649086\|2020- | ···················· | ···················· | ····················· | ···················· | ···················· | ···················· |

R 4837 hCoV-19/Israel/CVL-n3804/2020|EPI_ISL_649087|2020- ···················· ···················· ····················· ···················· ···················· ····················
R 4838 hCoV-19/Israel/CVL-n2971/2020|EPI_ISL_649088|2020- ···················· ···················· ····················· ···················· ···················· ····················
R 4839 hCoV-19/Israel/CVL-n3807/2020|EPI_ISL_649089|2020- ···················· ···················· ····················· ···················· ···················· ····················
R 4840 hCoV-19/Israel/CVL-n1467/2020|EPI_ISL_649090|2020- ···················· ···················· ····················· ···················· ···················· ····················
R 4841 hCoV-19/Israel/CVL-n2910/2020|EPI_ISL_649091|2020- ···················· ···················· ····················· ···················· ···················· ····················
R 4842 hCoV-19/Israel/CVL-n1999/2020|EPI_ISL_649092|2020- ···················· ···················· ····················· ···················· ···················· ····················
R 4843 hCoV-19/Israel/CVL-n2361/2020|EPI_ISL_649093|2020- ···················· ···················· ····················· ···················· ···················· ····················
R 4844 hCoV-19/Israel/CVL-n896/2020|EPI_ISL_649094|2020-0 ···················· ···················· ····················· ···················· ···················· ····················
R 4845 hCoV-19/Israel/CVL-n2200/2020|EPI_ISL_649095|2020- ···················· ···················· ····················· ···················· ···················· ····················
R 4846 hCoV-19/Israel/CVL-n2440/2020|EPI_ISL_649096|2020- ···················· ···················· ····················· ···················· ···················· ····················
R 4847 hCoV-19/Israel/CVL-n4094/2027|EPI_ISL_649097|2020- ···················· ···················· ····················· ···················· ···················· ····················
R 4848 hCoV-19/Israel/CVL-n1749/2020|EPI_ISL_649098|2020- ···················· ···················· ····················· ···················· ···················· ····················
R 4849 hCoV-19/Israel/CVL-n1750/2020|EPI_ISL_649099|2020- ···················· ···················· ····················· ···················· ···················· ····················
R 4850 hCoV-19/Israel/CVL-n2487/2020|EPI_ISL_649100|2020- ···················· ···················· ····················· ···················· ···················· ····················
R 4851 hCoV-19/Israel/CVL-n2775/2020|EPI_ISL_649101|2020- ···················· ···················· ····················· ···················· ···················· ····················
R 4852 hCoV-19/Israel/CVL-n3681/2020|EPI_ISL_649102|2020- ···················· ···················· ····················· ···················· ···················· ····················
R 4853 hCoV-19/Israel/CVL-n4431/2036|EPI_ISL_649103|2020- ···················· ···················· ····················· ···················· ···················· ····················
R 4854 hCoV-19/Israel/CVL-n3707/2020|EPI_ISL_649104|2020- ···················· ···················· ····················· ···················· ···················· ····················
R 4855 hCoV-19/Israel/CVL-n4065/2026|EPI_ISL_649105|2020- ···················· ···················· ····················· ···················· ···················· ····················
R 4856 hCoV-19/Israel/CVL-n4115/2031|EPI_ISL_649106|2020- ···················· ···················· ····················· ···················· ···················· ····················
R 4857 hCoV-19/Israel/CVL-n3975/2023|EPI_ISL_649107|2020- ···················· ···················· ····················· ···················· ···················· ····················
R 4858 hCoV-19/England/204721193/2020|EPI_ISL_649108|2020 ···················· ···················· ····················· ···················· ···················· ····················
R 4859 hCoV-19/England/204761593/2020|EPI_ISL_649112|2020 ···················· ···················· ····················· ···················· ···················· ····················
R 4860 hCoV-19/Australia/VIC16571/2020|EPI_ISL_649149|202 ···················· ···················· ····················· ···················· ···················· ····················
R 4861 hCoV-19/Australia/VIC12909/2020|EPI_ISL_649150|202 ···················· ···················· ····················· ···················· ···················· ····················
R 4862 hCoV-19/Australia/VIC16692/2020|EPI_ISL_649151|202 ···················· ···················· ····················· ···················· ···················· ····················
R 4863 hCoV-19/USA/CA-CZB-6808/2020|EPI_ISL_649152|2020-0 ···················· ·A·················· ····················· ···················· ···················· ····················
R 4864 hCoV-19/Palestine/WB-AQU-1/2020|EPI_ISL_649153|202 ···················· ···················· ····················· ···················· ···················· ····················
R 4865 hCoV-19/Australia/VIC16217/2020|EPI_ISL_640600|202 ···················· ···················· ····················· ···················· ···················· ····················
R 4866 hCoV-19/Australia/VIC16551/2020|EPI_ISL_640603|202 ···················· ···················· ····················· ···················· ···················· ····················
R 4867 hCoV-19/Australia/VIC16388/2020|EPI_ISL_640604|202 ···················· ···················· ····················· ···················· ···················· ····················
R 4868 hCoV-19/Australia/VIC16289/2020|EPI_ISL_640610|202 ···················· ···················· ····················· ···················· ···················· ····················
R 4869 hCoV-19/Australia/VIC12677/2020|EPI_ISL_640611|202 ···················· ···················· ····················· ···················· ···················· ····················
R 4870 hCoV-19/Australia/VIC16564/2020|EPI_ISL_640612|202 ···················· ···················· ····················· ···················· ···················· ····················
R 4871 hCoV-19/Australia/VIC11972/2020|EPI_ISL_640614|202 ···················· ···················· ····················· ···················· ···················· ····················
R 4872 hCoV-19/Australia/VIC12755/2020|EPI_ISL_640615|202 ···················· ···················· ····················· ···················· ···················· ····················
R 4873 hCoV-19/Australia/VIC16395/2020|EPI_ISL_640616|202 ···················· ···················· ····················· ···················· ···················· ····················
R 4874 hCoV-19/Australia/VIC11624/2020|EPI_ISL_640617|202 ···················· ···················· ····················· ···················· ···················· ····················
R 4875 hCoV-19/Australia/VIC12894/2020|EPI_ISL_640618|202 ···················· ···················· ····················· ···················· ···················· ····················
R 4876 hCoV-19/Australia/VIC16489/2020|EPI_ISL_640619|202 ···················· ···················· ····················· ···················· ···················· ····················
R 4877 hCoV-19/Australia/VIC13114/2020|EPI_ISL_640620|202 ···················· ···················· ····················· ···················· ···················· ····················
R 4878 hCoV-19/Australia/VIC16082/2020|EPI_ISL_640621|202 ···················· ···················· ····················· ···················· ···················· ····················
R 4879 hCoV-19/Australia/VIC12405/2020|EPI_ISL_640622|202 ···················· ···················· ····················· ···················· ···················· ····················
R 4880 hCoV-19/Australia/VIC11601/2020|EPI_ISL_640623|202 ···················· ···················· ····················· ···················· ···················· ····················
R 4881 hCoV-19/Australia/VIC12092/2020|EPI_ISL_640624|202 ···················· ···················· ····················· ···················· ···················· ····················
R 4884 hCoV-19/Australia/VIC16485/2020|EPI_ISL_640675|202 ···················· ···················· ····················· ···················· ···················· ····················
R 4885 hCoV-19/Australia/VIC12827/2020|EPI_ISL_640677|202 ···················· ···················· ····················· ···················· ···················· ····················
R 4886 hCoV-19/Australia/VIC12664/2020|EPI_ISL_640678|202 ···················· ···················· ····················· ···················· ···················· ····················
R 4887 hCoV-19/Australia/VIC12218/2020|EPI_ISL_640679|202 ···················· ···················· ····················· ···················· ···················· ····················
R 4888 hCoV-19/Australia/VIC12911/2020|EPI_ISL_640685|202 ···················· ···················· ····················· ···················· ···················· ····················
R 4889 hCoV-19/Australia/VIC12782/2020|EPI_ISL_640697|202 ···················· ···················· ····················· ···················· ···················· ····················
R 4890 hCoV-19/Australia/VIC12652/2020|EPI_ISL_640698|202 ···················· ···················· ····················· ···················· ···················· ····················
R 4891 hCoV-19/Australia/VIC12912/2020|EPI_ISL_640699|202 ···················· ···················· ····················· ···················· ···················· ····················
R 4892 hCoV-19/England/QEUH-B13D53/2020|EPI_ISL_641900|20 ···················· ···················· ····················· ···················· ···················· ····················
R 4893 hCoV-19/England/QEUH-B12D18/2020|EPI_ISL_641901|20 ···················· ···················· ····················· ···················· ···················· ····················
R 4894 hCoV-19/England/QEUH-B1301E/2020|EPI_ISL_641902|20 ···················· ···················· ····················· ···················· ···················· ····················
R 4895 hCoV-19/England/QEUH-B1345E/2020|EPI_ISL_641903|20 ···················· ···················· ····················· ···················· ···················· ····················
R 4896 hCoV-19/England/QEUH-B14262/2020|EPI_ISL_641904|20 ···················· ···················· ····················· ···················· ···················· ····················
R 4897 hCoV-19/England/QEUH-B142BD/2020|EPI_ISL_641905|20 ···················· ·A··G··············· ····················· ···················· ···················· ····················
R 4898 hCoV-19/England/QEUH-B140D1/2020|EPI_ISL_641906|20 ···················· ···················· ····················· ···················· ···················· ····················
R 4899 hCoV-19/England/QEUH-B13E9C/2020|EPI_ISL_641907|20 ···················· ·A·················· ····················· ···················· ···················· ····················
R 4900 hCoV-19/England/QEUH-B137A0/2020|EPI_ISL_641908|20 ···················· ···················· ····················· ···················· ···················· ····················
R 4901 hCoV-19/England/QEUH-B13CED/2020|EPI_ISL_641909|20 ···················· ···················· ····················· ···················· ···················· ····················
R 4902 hCoV-19/England/QEUH-B1349A/2020|EPI_ISL_641910|20 ···················· ···················· ····················· ···················· ······T············· ····················
R 4903 hCoV-19/England/QEUH-B137EC/2020|EPI_ISL_641911|20 ···················· ···················· ····················· ···················· ···················· ····················
R 4904 hCoV-19/England/QEUH-B0F75C/2020|EPI_ISL_641912|20 ···················· ·A·················· ····················· ···················· ···················· ····················
R 4905 hCoV-19/England/QEUH-B0E535/2020|EPI_ISL_641913|20 ···················· ···················· ····················· ···················· ···················· ····················
R 4906 hCoV-19/England/QEUH-B0EA90/2020|EPI_ISL_641914|20 ···················· ···················· ····················· ···················· ···················· ····················
R 4907 hCoV-19/England/QEUH-B0F367/2020|EPI_ISL_641915|20 ···················· ···················· ····················· ···················· ···················· ····················
R 4908 hCoV-19/England/QEUH-B110B6/2020|EPI_ISL_641916|20 ···················· ···················· ····················· ···················· ···················· ····················
R 4909 hCoV-19/England/QEUH-B0E15F/2020|EPI_ISL_641917|20 ···················· ···················· ····················· ···················· ···················· ····················
R 4910 hCoV-19/England/QEUH-B0F2D3/2020|EPI_ISL_641918|20 ···················· ···················· ····················· ···················· ···················· ····················
R 4911 hCoV-19/England/QEUH-B0E016/2020|EPI_ISL_641919|20 ···················· ···················· ····················· ···················· ···················· ····················
R 4912 hCoV-19/England/QEUH-B0F9B0/2020|EPI_ISL_641920|20 ···················· ···················· ····················· ···················· ···················· ····················
R 4913 hCoV-19/England/QEUH-B0EB24/2020|EPI_ISL_641921|20 ···················· ···················· ····················· ···················· ···················· ····················
R 4914 hCoV-19/England/QEUH-B0DB61/2020|EPI_ISL_641922|20 ···················· ···················· ····················· ···················· ···················· ····················
R 4915 hCoV-19/England/QEUH-B0F3D0/2020|EPI_ISL_641923|20 ···················· ···················· ····················· ···················· ···················· ····················
R 4916 hCoV-19/England/QEUH-B0DCAA/2020|EPI_ISL_641924|20 ···················· ···················· ····················· ···················· ···················· ····················
R 4917 hCoV-19/England/QEUH-B0E40B/2020|EPI_ISL_641925|20 ···················· ···················· ····················· ···················· ···················· ····················
R 4918 hCoV-19/England/QEUH-B0E8C3/2020|EPI_ISL_641926|20 ···················· ···················· ····················· ···················· ···················· ····················

R 4919 hCoV-19/England/QEUH-B0DDC5/2020|EPI_ISL_641927|20 .................... .................... ..................... .................... .................... ....................
R 4920 hCoV-19/England/QEUH-B0E632/2020|EPI_ISL_641928|20 .................... .................... ..................... .................... .................... ....................
R 4921 hCoV-19/England/QEUH-B0E2B6/2020|EPI_ISL_641929|20 .................... .................... ..................... .................... .................... ....................
R 4922 hCoV-19/England/QEUH-B0FCD5/2020|EPI_ISL_641930|20 .................... .A.................. ..................... .................... .................... ....................
R 4923 hCoV-19/England/QEUH-B0E359/2020|EPI_ISL_641931|20 .................... .................... ..................... .................... .................... ....................
R 4924 hCoV-19/England/QEUH-B0EA63/2020|EPI_ISL_641932|20 .................... .................... ..................... .................... .................... ....................
R 4925 hCoV-19/England/QEUH-B0E034/2020|EPI_ISL_641933|20 .................... .................... ..................... .................... .................... ....................
R 4926 hCoV-19/England/QEUH-B0E025/2020|EPI_ISL_641934|20 .................... .................... ..................... .................... .................... ....................
R 4927 hCoV-19/England/QEUH-B0EADC/2020|EPI_ISL_641935|20 .................... .A.................. ..................... .................... .................... ....................
R 4928 hCoV-19/England/QEUH-B0DA55/2020|EPI_ISL_641936|20 .................... .................... ..................... .................... .................... ...A................
R 4929 hCoV-19/England/QEUH-B0F956/2020|EPI_ISL_641937|20 .................... .................... ..................... .................... .................... ....................
R 4930 hCoV-19/England/QEUH-B0E4DE/2020|EPI_ISL_641938|20 .................... .................... ..................... .................... .................... ....................
R 4931 hCoV-19/England/QEUH-B0E3FF/2020|EPI_ISL_641939|20 .................... .................... ..................... .................... .................... ....................
R 4932 hCoV-19/England/QEUH-B0F2F1/2020|EPI_ISL_641941|20 .................... .................... ..................... .................... .................... ....................
R 4933 hCoV-19/England/QEUH-B0E571/2020|EPI_ISL_641942|20 .................... .................... ..................... .................... .................... ....................
R 4934 hCoV-19/England/QEUH-B0EA18/2020|EPI_ISL_641943|20 .................... .................... ..................... .................... .................... ....................
R 4935 hCoV-19/England/QEUH-B0EC4F/2020|EPI_ISL_641944|20 .................... .................... ..................... .................... .................... ....................
R 4936 hCoV-19/England/QEUH-B0E31D/2020|EPI_ISL_641945|20 .................... .................... ..................... .................... .................... ....................
R 4937 hCoV-19/England/QEUH-B0E508/2020|EPI_ISL_641946|20 .................... .A.................. ..................... .................... .................... ...A................
R 4938 hCoV-19/England/QEUH-B0DDE3/2020|EPI_ISL_641947|20 .................... .................... ..................... .................... .................... ....................
R 4939 hCoV-19/England/QEUH-B0E0DA/2020|EPI_ISL_641948|20 .................... .................... ..................... .................... .................... ....................
R 4940 hCoV-19/England/QEUH-B0E614/2020|EPI_ISL_641949|20 .................... .................... ..................... .................... .................... ....................
R 4941 hCoV-19/England/QEUH-B0EA45/2020|EPI_ISL_641951|20 .................... .................... ..................... .................... .................... ....................
R 4942 hCoV-19/England/QEUH-B0F40A/2020|EPI_ISL_641952|20 .................... .................... ..................... .................... .................... ....................
R 4943 hCoV-19/England/QEUH-B0E0AD/2020|EPI_ISL_641954|20 .................... .................... ..................... .................... .................... ....................
R 4944 hCoV-19/England/QEUH-B0E5AE/2020|EPI_ISL_641956|20 .................... .................... ..................... .................... .................... ....................
R 4945 hCoV-19/England/QEUH-B0FC02/2020|EPI_ISL_641957|20 .................... .................... ..................... .................... .................... ....................
R 4946 hCoV-19/England/QEUH-B0EB33/2020|EPI_ISL_641958|20 .................... .A.................. ..................... .................... .................... ....................
R 4947 hCoV-19/England/QEUH-B0E08F/2020|EPI_ISL_641959|20 .................... .................... ..................... .................... .................... ....................
R 4948 hCoV-19/England/QEUH-B0FB9C/2020|EPI_ISL_641961|20 .................... .................... ..................... .................... .................... ....................
R 4949 hCoV-19/England/204691653/2020|EPI_ISL_644162|2020 .................... .................... ..................... .................... .................... ....................
R 4950 hCoV-19/USA/TX-DSHS-1013/2020|EPI_ISL_644168|2020- .................... .................... ..................... .................... .................... ....................
R 4951 hCoV-19/USA/TX-DSHS-1022/2020|EPI_ISL_644176|2020- .................... .................... ..................... .................... .................... ....................
R 4952 hCoV-19/Ireland/D-NVRL-AIIDV1124v1/2020|EPI_ISL_64 .................... .................... ..................... .................... .................... ....................
R 4953 hCoV-19/Ireland/D-NVRL-AIIDM0045v1/2020|EPI_ISL_64 .................... .................... ..................... .................... .................... ....................
R 4954 hCoV-19/Ireland/D-NVRL-AIIDM0043v1/2020|EPI_ISL_64 .................... .................... ..................... .................... .................... ....................
R 4955 hCoV-19/Ireland/D-NVRL-AIIDM0017v1/2020|EPI_ISL_64 .................... .................... ..................... .................... .................... ....................
R 4956 hCoV-19/Ireland/D-NVRL-AIIDM0047v1/2020|EPI_ISL_64 .................... .................... ..................... .................... .................... ....................
R 4957 hCoV-19/Ireland/D-NVRL-AIIDM0040v1/2020|EPI_ISL_64 .................... .................... ..................... .................... .................... ....................
R 4958 hCoV-19/Ireland/WW-NVRL-AIIDV0379v2/2020|EPI_ISL_6 .................... .................... ..................... .................... .................... ....................
R 4959 hCoV-19/Ireland/MH-NVRL-AIIDM0073v1/2020|EPI_ISL_6 .................... .................... ..................... .................... .................... ....................
R 4960 hCoV-19/Ireland/D-NVRL-AIIDV0072v2/2020|EPI_ISL_64 .................... .................... ..................... .................... .................... ....................
R 4961 hCoV-19/Ireland/D-NVRL-AIIDM0079v1/2020|EPI_ISL_64 .................... .................... ..................... .................... .................... ....................
R 4962 hCoV-19/Ireland/WW-NVRL-AIIDV0063v2/2020|EPI_ISL_6 .................... .................... ..................... .................... .................... ....................
R 4963 hCoV-19/Ireland/D-NVRL-AIIDV0066v1/2020|EPI_ISL_64 .................... .................... ..................... .................... .................... ....................
R 4964 hCoV-19/Ireland/D-NVRL-AIIDV0125v1/2020|EPI_ISL_64 .................... .................... ..................... .................... .................... ....................
R 4965 hCoV-19/Ireland/D-NVRL-AIIDV0103v1/2020|EPI_ISL_64 .................... .................... ..................... .................... .................... ....................
R 4966 hCoV-19/Ireland/D-NVRL-AIIDV1131v1/2020|EPI_ISL_64 .................... .................... ..................... .................... .................... ....................
R 4967 hCoV-19/Ireland/WW-NVRL-AIIDV1109v1/2020|EPI_ISL_6 .................... .................... ..................... .................... .................... ....................
R 4968 hCoV-19/Ireland/D-NVRL-AIIDV0004v1/2020|EPI_ISL_64 .................... .................... ..................... .................... .................... ....................
R 4969 hCoV-19/Ireland/D-NVRL-AIIDV0106v1/2020|EPI_ISL_64 .................... .................... ..................... .................... .................... ....................
R 4970 hCoV-19/Ireland/D-NVRL-AIIDV0023v1/2020|EPI_ISL_64 .................... .................... ..................... .................... .................... ....................
R 4971 hCoV-19/Ireland/D-NVRL-AIIDV0021v1/2020|EPI_ISL_64 .................... .................... ..................... .................... .................... ....................
R 4972 hCoV-19/Ireland/CE-NVRL-AIIDV1105v2/2020|EPI_ISL_6 .................... .................... ..................... .................... .................... ....................
R 4973 hCoV-19/Ireland/WW-NVRL-AIIDV0073v1/2020|EPI_ISL_6 .................... .................... ..................... .................... .................... ....................
R 4974 hCoV-19/Ireland/D-NVRL-AIIDV0080v1/2020|EPI_ISL_64 .................... .................... ..................... .................... .................... ....................
R 4975 hCoV-19/Ireland/CE-NVRL-AIIDV1105v4/2020|EPI_ISL_6 .................... .................... ..................... .................... .................... ....................
R 4977 hCoV-19/Ireland/D-NVRL-AIIDV0053v1/2020|EPI_ISL_64 .................... .................... ..................... .................... .................... ....................
R 4978 hCoV-19/Ireland/LD-NVRL-AIIDV0197v1/2020|EPI_ISL_6 .................... .................... ................G.... .................... .................... ....................
R 4979 hCoV-19/Ireland/D-NVRL-AIIDV0072v1/2020|EPI_ISL_64 .................... .................... ..................... .................... .................... ....................
R 4980 hCoV-19/Ireland/WW-NVRL-AIIDV0073v2/2020|EPI_ISL_6 .................... .................... ..................... .................... .................... ....................
R 4982 hCoV-19/Ireland/D-NVRL-AIIDV0081v1/2020|EPI_ISL_64 .................... .................... ..................... .................... .................... ....................
R 4983 hCoV-19/England/CAMC-B35E49/2020|EPI_ISL_645492|20 .................... .................... ..................... .................... .................... ....................
R 4984 hCoV-19/England/CAMC-B364DD/2020|EPI_ISL_645495|20 .................... .................... ..................... .................... .................... ....................
R 4985 hCoV-19/England/CAMC-B363D0/2020|EPI_ISL_645496|20 .................... .................... ..................... .................... ......T............. ....................
R 4986 hCoV-19/England/CAMC-B37339/2020|EPI_ISL_645497|20 .................... .................... ..................... .................... .................... ...A................
R 4987 hCoV-19/England/CAMC-B3706F/2020|EPI_ISL_645498|20 .................... .................... ..................... .................... .................... ....................
R 4988 hCoV-19/England/ALDP-B3E55A/2020|EPI_ISL_645499|20 .................... .................... ..................... .................... .................... ....................
R 4989 hCoV-19/England/CAMC-B364CE/2020|EPI_ISL_645526|20 .................... .................... ..................... .................... .................... ....................
R 4990 hCoV-19/England/CAMC-B372F0/2020|EPI_ISL_645528|20 .................... .................... ..................... .................... .................... ....................
R 4991 hCoV-19/England/CAMC-B3715D/2020|EPI_ISL_645529|20 .................... .................... ..................... .................... .................... ....................
R 4992 hCoV-19/England/ALDP-B1BB60/2020|EPI_ISL_645530|20 .................... .................... ..................... .................... .................... ....................
R 4993 hCoV-19/England/ALDP-B1AF65/2020|EPI_ISL_645531|20 .................... .................... ..................... .................... .................... ....................
R 4994 hCoV-19/England/ALDP-B1B2F2/2020|EPI_ISL_645532|20 .................... .................... ..................... .................... .................... ....................
R 4995 hCoV-19/England/ALDP-B1B40B/2020|EPI_ISL_645533|20 .................... .................... ..................... .................... .................... ....................
R 4996 hCoV-19/England/ALDP-B1B3A4/2020|EPI_ISL_645534|20 .................... .................... ..................... .................... .................... ....................
R 4997 hCoV-19/England/ALDP-B1CDA5/2020|EPI_ISL_645535|20 .................... .................... ..................... .................... .................... ....................
R 4998 hCoV-19/England/ALDP-B1CD96/2020|EPI_ISL_645537|20 .................... .................... ..................... .................... .................... ....................
R 4999 hCoV-19/England/ALDP-B1A080/2020|EPI_ISL_645538|20 .................... .................... ..................... .................... .................... ....................
R 5000 hCoV-19/England/ALDP-B1A16F/2020|EPI_ISL_645539|20 .................... .A.................. ..................... .................... .................... ....................

| | | | | | | | | |
|---|---|---|---|---|---|---|---|---|
| R | 5001 | hCoV-19/England/ALDP-B1BF82/2020\|EPI_ISL_645540\|20 | ···················· | ···················· | ····················· | ···················· | ···················· | ···················· |
| R | 5002 | hCoV-19/England/ALDP-B1CE2A/2020\|EPI_ISL_645541\|20 | ···················· | ···················· | ····················· | ···················· | ···················· | ···················· |
| R | 5003 | hCoV-19/England/ALDP-B1CEEE/2020\|EPI_ISL_645542\|20 | ···················· | ···················· | ····················· | ···················· | ···················· | ···················· |
| R | 5004 | hCoV-19/England/ALDP-B1BB15/2020\|EPI_ISL_645543\|20 | ···················· | ···················· | ····················· | ···················· | ···················· | ···················· |
| R | 5005 | hCoV-19/England/ALDP-B1CE66/2020\|EPI_ISL_645544\|20 | ···················· | ···················· | ····················· | ···················· | ···················· | ···················· |
| R | 5006 | hCoV-19/England/ALDP-B1A17E/2020\|EPI_ISL_645545\|20 | ···················· | ···················· | ····················· | ···················· | ···················· | ···················· |
| R | 5007 | hCoV-19/England/ALDP-B1B993/2020\|EPI_ISL_645546\|20 | ···················· | ···················· | ····················· | ···················· | ···················· | ···················· |
| R | 5008 | hCoV-19/England/ALDP-B1E2B3/2020\|EPI_ISL_645548\|20 | ···················· | ···················· | ····················· | ···················· | ···················· | ···················· |
| R | 5009 | hCoV-19/England/ALDP-B1C561/2020\|EPI_ISL_645549\|20 | ···················· | ···················· | ····················· | ···················· | ···················· | ···················· |
| R | 5010 | hCoV-19/England/ALDP-B1A633/2020\|EPI_ISL_645550\|20 | ···················· | ···················· | ····················· | ···················· | ···················· | ···················· |
| R | 5011 | hCoV-19/England/ALDP-B1A19C/2020\|EPI_ISL_645551\|20 | ···················· | ···················· | ····················· | ···················· | ···················· | ···················· |
| R | 5012 | hCoV-19/England/ALDP-B1B016/2020\|EPI_ISL_645552\|20 | ···················· | ···················· | ····················· | ···················· | ···················· | ···················· |
| R | 5013 | hCoV-19/England/ALDP-B1BFFB/2020\|EPI_ISL_645553\|20 | ···················· | ···················· | ····················· | ···················· | ···················· | ···················· |
| R | 5014 | hCoV-19/England/ALDP-B1B492/2020\|EPI_ISL_645555\|20 | ···················· | ···················· | ····················· | ···················· | ···················· | ···················· |
| R | 5015 | hCoV-19/England/ALDP-B1B0DA/2020\|EPI_ISL_645556\|20 | ···················· | ···················· | ···A················· | ···················· | ···················· | ···················· |
| R | 5016 | hCoV-19/England/ALDP-B1B9B1/2020\|EPI_ISL_645557\|20 | ···················· | ···················· | ····················· | ···················· | ···················· | ···················· |
| R | 5017 | hCoV-19/England/ALDP-B1B5EA/2020\|EPI_ISL_645558\|20 | ···················· | ···················· | ····················· | ···················· | ···················· | ···················· |
| R | 5018 | hCoV-19/England/ALDP-B1B3B3/2020\|EPI_ISL_645559\|20 | ···················· | ···················· | ····················· | ···················· | ···················· | ···················· |
| R | 5019 | hCoV-19/England/ALDP-B1A518/2020\|EPI_ISL_645560\|20 | ···················· | ···················· | ····················· | ···················· | ···················· | ···················· |
| R | 5020 | hCoV-19/England/ALDP-B1CE0C/2020\|EPI_ISL_645561\|20 | ···················· | ···················· | ····················· | ···················· | ···················· | ···················· |
| R | 5021 | hCoV-19/England/ALDP-B1B395/2020\|EPI_ISL_645562\|20 | ···················· | ···················· | ····················· | ···················· | ···················· | ···················· |
| R | 5022 | hCoV-19/England/ALDP-B1BAAF/2020\|EPI_ISL_645563\|20 | ···················· | ···················· | ····················· | ···················· | ···················· | ···················· |
| R | 5023 | hCoV-19/England/ALDP-B1B447/2020\|EPI_ISL_645564\|20 | ···················· | ···················· | ····················· | ···················· | ···················· | ···················· |
| R | 5024 | hCoV-19/England/ALDP-B1B3FF/2020\|EPI_ISL_645565\|20 | ···················· | ···················· | ····················· | ···················· | ···················· | ···················· |
| R | 5025 | hCoV-19/England/ALDP-B1B5BD/2020\|EPI_ISL_645566\|20 | ···················· | ···················· | ····················· | ···················· | ···················· | ···················· |
| R | 5026 | hCoV-19/England/ALDP-B1B966/2020\|EPI_ISL_645567\|20 | ···················· | ···················· | ····················· | ···················· | ···················· | ···················· |
| R | 5027 | hCoV-19/England/ALDP-B1BB9D/2020\|EPI_ISL_645568\|20 | ···················· | ···················· | ····················· | ···················· | ···················· | ···················· |
| R | 5028 | hCoV-19/England/ALDP-B1BBBB/2020\|EPI_ISL_645569\|20 | ···················· | ···················· | ····················· | ···················· | ···················· | ···················· |
| R | 5029 | hCoV-19/England/ALDP-B1ABF8/2020\|EPI_ISL_645570\|20 | ···················· | ···················· | ····················· | ···················· | ···················· | ···················· |
| R | 5030 | hCoV-19/England/ALDP-B1BC30/2020\|EPI_ISL_645571\|20 | ···················· | ···················· | ····················· | ···················· | ···················· | ···················· |
| R | 5031 | hCoV-19/England/ALDP-B1A132/2020\|EPI_ISL_645572\|20 | ···················· | ·A·················· | ····················· | ···················· | ···················· | ···················· |
| R | 5032 | hCoV-19/England/ALDP-B1D050/2020\|EPI_ISL_645573\|20 | ···················· | ···················· | ····················· | ···················· | ···················· | ···················· |
| R | 5033 | hCoV-19/England/ALDP-B1B562/2020\|EPI_ISL_645574\|20 | ···················· | ···················· | ····················· | ···················· | ···················· | ···················· |
| R | 5034 | hCoV-19/England/ALDP-B1BF55/2020\|EPI_ISL_645575\|20 | ···················· | ···················· | ····················· | ···················· | ···················· | ···················· |
| R | 5035 | hCoV-19/England/ALDP-B1A09F/2020\|EPI_ISL_645576\|20 | ···················· | ···················· | ····················· | ···················· | ···················· | ···················· |
| R | 5036 | hCoV-19/England/ALDP-B1BB42/2020\|EPI_ISL_645577\|20 | ···················· | ···················· | ····················· | ···················· | ···················· | ···················· |
| R | 5037 | hCoV-19/England/ALDP-B1BBD9/2020\|EPI_ISL_645578\|20 | ···················· | ···················· | ····················· | ···················· | ···················· | ···················· |
| R | 5038 | hCoV-19/England/ALDP-B1CCF3/2020\|EPI_ISL_645579\|20 | ···················· | ···················· | ····················· | ···················· | ···················· | ···················· |
| R | 5039 | hCoV-19/England/ALDP-B1B508/2020\|EPI_ISL_645580\|20 | ···················· | ···················· | ····················· | ···················· | ···················· | ···················· |
| R | 5040 | hCoV-19/England/ALDP-B1BF64/2020\|EPI_ISL_645581\|20 | ···················· | ···················· | ····················· | ···················· | ···················· | ···················· |
| R | 5041 | hCoV-19/England/ALDP-B1A79A/2020\|EPI_ISL_645582\|20 | ···················· | ···················· | ····················· | ···················· | ···················· | ···················· |
| R | 5042 | hCoV-19/England/ALDP-B1A123/2020\|EPI_ISL_645584\|20 | ···················· | ···················· | ····················· | ···················· | ···················· | ···················· |
| R | 5043 | hCoV-19/England/ALDP-B1B5CC/2020\|EPI_ISL_645585\|20 | ···················· | ···················· | ····················· | ···················· | ······T············· | ···················· |
| R | 5044 | hCoV-19/England/ALDP-B1C4FB/2020\|EPI_ISL_645586\|20 | ···················· | ···················· | ····················· | ···················· | ···················· | ···················· |
| R | 5045 | hCoV-19/England/ALDP-B1B025/2020\|EPI_ISL_645587\|20 | ···················· | ···················· | ····················· | ···················· | ···················· | ···················· |
| R | 5046 | hCoV-19/England/ALDP-B1C349/2020\|EPI_ISL_645588\|20 | ···················· | ···················· | ····················· | ···················· | ···················· | ···················· |
| R | 5047 | hCoV-19/England/ALDP-B1B623/2020\|EPI_ISL_645589\|20 | ···················· | ···················· | ····················· | ···················· | ···················· | ···················· |
| R | 5048 | hCoV-19/England/ALDP-B1C428/2020\|EPI_ISL_645590\|20 | ···················· | ···················· | ····················· | ···················· | ······T············· | ···················· |
| R | 5049 | hCoV-19/England/ALDP-B1CEC0/2020\|EPI_ISL_645591\|20 | ···················· | ···················· | ····················· | ···················· | ···················· | ···················· |
| R | 5050 | hCoV-19/England/ALDP-B1B4B0/2020\|EPI_ISL_645592\|20 | ···················· | ·········T·········· | ····················· | ···················· | ···················· | ···················· |
| R | 5051 | hCoV-19/England/ALDP-B1B957/2020\|EPI_ISL_645593\|20 | ···················· | ···················· | ····················· | ···················· | ···················· | ···················· |
| R | 5052 | hCoV-19/England/ALDP-B1AF47/2020\|EPI_ISL_645594\|20 | ···················· | ···················· | ····················· | ···················· | ···················· | ···················· |
| R | 5053 | hCoV-19/England/ALDP-B1B5F9/2020\|EPI_ISL_645595\|20 | ···················· | ···················· | ····················· | ···················· | ···················· | ···················· |
| R | 5054 | hCoV-19/England/ALDP-B1BF91/2020\|EPI_ISL_645596\|20 | ···················· | ···················· | ····················· | ···················· | ···················· | ···················· |
| R | 5055 | hCoV-19/England/ALDP-B1BB06/2020\|EPI_ISL_645597\|20 | ···················· | ···················· | ····················· | ···················· | ···················· | ···················· |
| R | 5056 | hCoV-19/England/ALDP-B1B9EE/2020\|EPI_ISL_645598\|20 | ···················· | ···················· | ····················· | ···················· | ···················· | ···················· |
| R | 5057 | hCoV-19/England/ALDP-B1BABE/2020\|EPI_ISL_645599\|20 | ···················· | ···················· | ····················· | ···················· | ···················· | ···················· |
| R | 5058 | hCoV-19/England/ALDP-B24715/2020\|EPI_ISL_646857\|20 | ···················· | ···················· | ····················· | ···················· | ···················· | ···················· |
| R | 5059 | hCoV-19/England/ALDP-B23A59/2020\|EPI_ISL_646858\|20 | ···················· | ···················· | ····················· | ···················· | ···················· | ···················· |
| R | 5060 | hCoV-19/England/ALDP-B2440F/2020\|EPI_ISL_646859\|20 | ···················· | ···················· | ····················· | ···················· | ···················· | ···················· |
| R | 5061 | hCoV-19/England/ALDP-B24320/2020\|EPI_ISL_646860\|20 | ···················· | ···················· | ····················· | ···················· | ···················· | ···················· |
| R | 5062 | hCoV-19/England/ALDP-B23AD1/2020\|EPI_ISL_646861\|20 | ···················· | ···················· | ····················· | ···················· | ···················· | ···················· |
| R | 5063 | hCoV-19/England/ALDP-B23822/2020\|EPI_ISL_646862\|20 | ···················· | ···················· | ····················· | ···················· | ···················· | ···················· |
| R | 5064 | hCoV-19/England/ALDP-B2379E/2020\|EPI_ISL_646863\|20 | ···················· | ···················· | ····················· | ···················· | ···················· | ···················· |
| R | 5065 | hCoV-19/England/ALDP-B24706/2020\|EPI_ISL_646864\|20 | ···················· | ···················· | ····················· | ···················· | ···················· | ···················· |
| R | 5066 | hCoV-19/England/ALDP-B27448/2020\|EPI_ISL_646865\|20 | ···················· | ···················· | ····················· | ···················· | ···················· | ···················· |
| R | 5067 | hCoV-19/England/ALDP-B2450C/2020\|EPI_ISL_646866\|20 | ···················· | ···················· | ····················· | ···················· | ···················· | ···················· |
| R | 5068 | hCoV-19/England/ALDP-B24126/2020\|EPI_ISL_646867\|20 | ···················· | ·A·················· | ····················· | ···················· | ···················· | ···················· |
| R | 5069 | hCoV-19/England/QEUH-B2DB4D/2020\|EPI_ISL_646868\|20 | ···················· | ···················· | ····················· | ···················· | ···················· | ···················· |
| R | 5070 | hCoV-19/England/QEUH-B2D758/2020\|EPI_ISL_646869\|20 | ···················· | ···················· | ····················· | ···················· | ···················· | ···················· |
| R | 5071 | hCoV-19/England/ALDP-B24803/2020\|EPI_ISL_646871\|20 | ···················· | ···················· | ····················· | ···················· | ···················· | ···················· |
| R | 5072 | hCoV-19/England/QEUH-B2EC76/2020\|EPI_ISL_646872\|20 | ···················· | ···················· | ····················· | ···················· | ···················· | ···················· |
| R | 5073 | hCoV-19/England/ALDP-B248F4/2020\|EPI_ISL_646873\|20 | ···················· | ···················· | ····················· | ···················· | ···················· | ···················· |
| R | 5074 | hCoV-19/England/QEUH-B2DC3B/2020\|EPI_ISL_646874\|20 | ···················· | ···················· | ····················· | ···················· | ···················· | ···················· |
| R | 5075 | hCoV-19/England/ALDP-B248B8/2020\|EPI_ISL_646875\|20 | ···················· | ···················· | ····················· | ···················· | ···················· | ···················· |
| R | 5076 | hCoV-19/England/ALDP-B24487/2020\|EPI_ISL_646876\|20 | ···················· | ···················· | ····················· | ···················· | ···················· | ···················· |
| R | 5077 | hCoV-19/England/QEUH-B2DA22/2020\|EPI_ISL_646877\|20 | ···················· | ···················· | ····················· | ···················· | ···················· | ···················· |
| R | 5078 | hCoV-19/England/ALDP-B2494C/2020\|EPI_ISL_646878\|20 | ···················· | ···················· | ····················· | ···················· | ···················· | ···················· |
| R | 5079 | hCoV-19/England/QEUH-B2D6E2/2020\|EPI_ISL_646880\|20 | ···················· | ·A·················· | ····················· | ···················· | ···················· | ···A················ |
| R | 5080 | hCoV-19/England/ALDP-B23840/2020\|EPI_ISL_646881\|20 | ···················· | ···················· | ····················· | ···················· | ···················· | ···················· |

```
R 5081 hCoV-19/England/ALDP-B2444B/2020|EPI_ISL_646883|20   ···················· ···················· ···················· ···················· ···················· ····················
R 5082 hCoV-19/England/ALDP-B24AEF/2020|EPI_ISL_646884|20   ···················· ···················· ···················· ···················· ···················· ····················
R 5083 hCoV-19/England/QEUH-B2DB01/2020|EPI_ISL_646885|20   ···················· ···················· ···················· ···················· ···················· ····················
R 5084 hCoV-19/England/ALDP-B23655/2020|EPI_ISL_646886|20   ···················· ···················· ···················· ···················· ···················· ····················
R 5085 hCoV-19/England/ALDP-B237DA/2020|EPI_ISL_646887|20   ···················· ···················· ···················· ···················· ···················· ····················
R 5086 hCoV-19/England/ALDP-B23A0E/2020|EPI_ISL_646888|20   ···················· ···················· ···················· ···················· ···················· ····················
R 5087 hCoV-19/England/ALDP-B27439/2020|EPI_ISL_646889|20   ···················· ···················· ···················· ···················· ······T············· ····················
R 5088 hCoV-19/England/ALDP-B244E1/2020|EPI_ISL_646890|20   ···················· ···················· ···················· ···················· ···················· ····················
R 5089 hCoV-19/England/QEUH-B2EC85/2020|EPI_ISL_646891|20   ···················· ···················· ···················· ···················· ···················· ····················
R 5090 hCoV-19/England/QEUH-B2DB3E/2020|EPI_ISL_646892|20   ···················· ···················· ···················· ···················· ···················· ····················
R 5091 hCoV-19/England/ALDP-B24469/2020|EPI_ISL_646893|20   ···················· ···················· ···················· ···················· ···················· ····················
R 5092 hCoV-19/England/ALDP-B2388C/2020|EPI_ISL_646894|20   ···················· ···················· ···················· ···················· ···················· ····················
R 5093 hCoV-19/England/ALDP-B2378F/2020|EPI_ISL_646895|20   ···················· ···················· ···················· ···················· ···················· ····················
R 5094 hCoV-19/England/ALDP-B24B73/2020|EPI_ISL_646896|20   ···················· ···················· ···················· ···················· ···················· ····················
R 5095 hCoV-19/England/ALDP-B2436C/2020|EPI_ISL_646897|20   ···················· ···················· ···················· ···················· ···················· ····················
R 5096 hCoV-19/England/ALDP-B247D9/2020|EPI_ISL_646898|20   ···················· ··············C····· ···················· ···················· ···················· ····················
R 5097 hCoV-19/England/ALDP-B236FB/2020|EPI_ISL_646899|20   ···················· ···················· ···················· ···················· ···················· ····················
R 5098 hCoV-19/USA/CA-ALSR-4404/2020|EPI_ISL_649010|2020-   ···················· ···················· ···················· ···················· ···················· ····················
R 5099 hCoV-19/USA/CA-ALSR-4405/2020|EPI_ISL_649011|2020-   ···················· ·A·················· ···················· ···················· ···················· ····················
R 5100 hCoV-19/USA/CA-ALSR-4407/2020|EPI_ISL_649012|2020-   ···················· ···················· ···················· ···················· ···················· ····················
R 5101 hCoV-19/USA/CA-ALSR-4409/2020|EPI_ISL_649013|2020-   ···················· ·A·················· ···················· ···················· ···················· ····················
R 5102 hCoV-19/USA/CA-ALSR-4411/2020|EPI_ISL_649014|2020-   ···················· ···················· ···················· ···················· ···················· ····················
R 5103 hCoV-19/USA/CA-ALSR-4437/2020|EPI_ISL_649030|2020-   ···················· ···················· ···················· ···················· ···················· ····················
R 5104 hCoV-19/USA/CA-ALSR-4440/2020|EPI_ISL_649032|2020-   ···················· ·A·················· ···················· ···················· ···················· ····················
R 5105 hCoV-19/USA/CA-ALSR-4442/2020|EPI_ISL_649033|2020-   ···················· ·A·················· ···················· ···················· ···················· ····················
R 5106 hCoV-19/USA/CA-ALSR-4447/2020|EPI_ISL_649034|2020-   ···················· ·A·················· ···················· ···················· ···················· ····················
R 5107 hCoV-19/USA/CA-ALSR-4450/2020|EPI_ISL_649036|2020-   ···················· ···················· ···················· ···················· ···················· ····················
R 5108 hCoV-19/USA/CA-ALSR-4451/2020|EPI_ISL_649037|2020-   ···················· ·A·················· ···················· ···················· ···················· ····················
R 5109 hCoV-19/USA/CA-ALSR-4452/2020|EPI_ISL_649038|2020-   ···················· ·A·················· ···················· ···················· ···················· ····················
R 5110 hCoV-19/USA/CA-ALSR-4455/2020|EPI_ISL_649040|2020-   ···················· ·A·················· ···················· ···················· ···················· ····················
R 5111 hCoV-19/USA/CA-ALSR-4459/2020|EPI_ISL_649042|2020-   ···················· ·A·················· ···················· ···················· ···················· ····················
R 5112 hCoV-19/USA/CA-ALSR-4461/2020|EPI_ISL_649043|2020-   ···················· ···················· ···················· ···················· ···················· ····················
R 5113 hCoV-19/USA/CA-ALSR-4462/2020|EPI_ISL_649044|2020-   ···················· ·A·················· ···················· ···················· ···················· ····················
R 5114 hCoV-19/USA/CA-ALSR-4463/2020|EPI_ISL_649045|2020-   ···················· ·A·················· ···················· ···················· ···················· ····················
R 5115 hCoV-19/USA/CA-ALSR-4464/2020|EPI_ISL_649046|2020-   ···················· ·A·················· ···················· ···················· ···················· ····················
R 5116 hCoV-19/USA/CA-ALSR-4466/2020|EPI_ISL_649047|2020-   ···················· ·A·················· ···················· ···················· ···················· ····················
R 5117 hCoV-19/USA/CA-ALSR-4468/2020|EPI_ISL_649048|2020-   ···················· ·A·················· ···················· ···················· ···················· ····················
R 5118 hCoV-19/USA/CA-ALSR-4472/2020|EPI_ISL_649049|2020-   ···················· ·A·················· ···················· ···················· ···················· ····················
R 5119 hCoV-19/Israel/CVL-n1541/2020|EPI_ISL_649064|2020-   ···················· ···················· ···················· ···················· ···················· ····················
R 5120 hCoV-19/Israel/CVL-n3782/2020|EPI_ISL_649065|2020-   ···················· ···················· ···················· ···················· ···················· ····················
R 5121 hCoV-19/Australia/VIC12609/2020|EPI_ISL_640598|202   ···················· ···················· ···················· ···················· ···················· ····················
R 5122 hCoV-19/Australia/VIC16158/2020|EPI_ISL_640631|202   ···················· ···················· ···················· ···················· ···················· ····················
R 5123 hCoV-19/Australia/VIC16311/2020|EPI_ISL_640632|202   ···················· ···················· ···················· ···················· ···················· ····················
R 5124 hCoV-19/Australia/VIC15960/2020|EPI_ISL_640633|202   ···················· ···················· ···················· ···················· ···················· ····················
R 5125 hCoV-19/Australia/VIC12147/2020|EPI_ISL_640635|202   ···················· ···················· ···················· ···················· ···················· ····················
R 5126 hCoV-19/Australia/VIC16409/2020|EPI_ISL_640636|202   ···················· ···················· ···················· ···················· ···················· ····················
R 5128 hCoV-19/Australia/VIC16103/2020|EPI_ISL_640638|202   ···················· ···················· ···················· ···················· ···················· ····················
R 5129 hCoV-19/Australia/VIC16416/2020|EPI_ISL_640642|202   ···················· ···················· ···················· ···················· ···················· ····················
R 5131 hCoV-19/Australia/VIC12702/2020|EPI_ISL_640646|202   ···················· ···················· ···················· ···················· ···················· ····················
R 5132 hCoV-19/Australia/VIC15942/2020|EPI_ISL_640647|202   ···················· ···················· ···················· ···················· ···················· ····················
R 5133 hCoV-19/Australia/VIC16051/2020|EPI_ISL_640648|202   ···················· ···················· ···················· ···················· ···················· ····················
R 5134 hCoV-19/Australia/VIC16341/2020|EPI_ISL_640649|202   ···················· ···················· ···················· ···················· ···················· ····················
R 5136 hCoV-19/Australia/VIC15933/2020|EPI_ISL_640652|202   ···················· ···················· ···················· ···················· ···················· ····················
R 5137 hCoV-19/Australia/VIC16092/2020|EPI_ISL_640653|202   ···················· ···················· ···················· ···················· ···················· ····················
R 5138 hCoV-19/Australia/VIC12842/2020|EPI_ISL_640655|202   ···················· ······A············· ···················· ···················· ···················· ····················
R 5139 hCoV-19/Australia/VIC12629/2020|EPI_ISL_640656|202   ···················· ···················· ···················· ···················· ···················· ····················
R 5140 hCoV-19/Australia/VIC16191/2020|EPI_ISL_640657|202   ···················· ···················· ···················· ···················· ···················· ····················
R 5141 hCoV-19/Australia/VIC12425/2020|EPI_ISL_640659|202   ···················· ···················· ···················· ···················· ···················· ····················
R 5143 hCoV-19/Australia/VIC12581/2020|EPI_ISL_640661|202   ···················· ···················· ···················· ···················· ···················· ····················
R 5145 hCoV-19/Australia/VIC12241/2020|EPI_ISL_640663|202   ···················· ···················· ···················· ···················· ···················· ····················
R 5146 hCoV-19/Australia/VIC16243/2020|EPI_ISL_640665|202   ···················· ···················· ···················· ···················· ···················· ····················
R 5148 hCoV-19/Australia/VIC11631/2020|EPI_ISL_640667|202   ···················· ···················· ···················· ···················· ···················· ····················
R 5149 hCoV-19/Australia/VIC12463/2020|EPI_ISL_640668|202   ···················· ···················· ···················· ···················· ···················· ····················
R 5151 hCoV-19/Australia/VIC16436/2020|EPI_ISL_640671|202   ···················· ···················· ···················· ···················· ···················· ····················
R 5152 hCoV-19/Australia/VIC12598/2020|EPI_ISL_640672|202   ···················· ···················· ···················· ···················· ···················· ····················
R 5153 hCoV-19/Australia/VIC16498/2020|EPI_ISL_640673|202   ···················· ···················· ···················· ···················· ···················· ····················
R 5154 hCoV-19/Australia/VIC15939/2020|EPI_ISL_640684|202   ···················· ···················· ···················· ···················· ···················· ····················
R 5155 hCoV-19/Australia/VIC16015/2020|EPI_ISL_640686|202   ···················· ···················· ···················· ···················· ···················· ····················
R 5156 hCoV-19/Australia/VIC15916/2020|EPI_ISL_640687|202   ···················· ···················· ···················· ···················· ···················· ····················
R 5157 hCoV-19/Australia/VIC12544/2020|EPI_ISL_640688|202   ···················· ···················· ···················· ···················· ···················· ····················
R 5160 hCoV-19/Australia/VIC12703/2020|EPI_ISL_640691|202   ···················· ···················· ···················· ···················· ···················· ····················
R 5161 hCoV-19/Australia/VIC16628/2020|EPI_ISL_640692|202   ···················· ···················· ···················· ···················· ···················· ····················
R 5163 hCoV-19/Australia/VIC12889/2020|EPI_ISL_640694|202   ···················· ···················· ···················· ···················· ···················· ····················
R 5165 hCoV-19/England/QEUH-B0DB8F/2020|EPI_ISL_641962|20   ···················· ···················· ···················· ···················· ···················· ····················
R 5166 hCoV-19/England/QEUH-B0F895/2020|EPI_ISL_641963|20   ···················· ···················· ···················· ···················· ···················· ····················
R 5167 hCoV-19/England/QEUH-B0E1AA/2020|EPI_ISL_641964|20   ···················· ···················· ···················· ···················· ···················· ····················
R 5168 hCoV-19/England/QEUH-B0DD10/2020|EPI_ISL_641965|20   ···················· ···················· ···················· ···················· ···················· ····················
R 5169 hCoV-19/England/QEUH-B0F419/2020|EPI_ISL_641966|20   ···················· ···················· ···················· ···················· ···················· ····················
R 5170 hCoV-19/England/QEUH-B0F376/2020|EPI_ISL_641967|20   ···················· ···················· ···················· ···················· ···················· ····················
R 5171 hCoV-19/England/QEUH-B0E8A5/2020|EPI_ISL_641968|20   ···················· ···················· ···················· ···················· ···················· ····················
```

| | | | | | | | |
|---|---|---|---|---|---|---|---|
| R | 5172 hCoV-19/England/QEUH-B0DA73/2020\|EPI_ISL_641969\|20 | ···················· | ···················· | ···················· | ···················· | ···················· | ···················· |
| R | 5173 hCoV-19/England/QEUH-B0F2C4/2020\|EPI_ISL_641970\|20 | ···················· | ···················· | ···················· | ···················· | ····T··············· | ···················· |
| R | 5174 hCoV-19/England/QEUH-B0FC20/2020\|EPI_ISL_641971\|20 | ···················· | ···················· | ···················· | ···················· | ···················· | ···················· |
| R | 5175 hCoV-19/England/QEUH-B0DEE0/2020\|EPI_ISL_641972\|20 | ···················· | ···················· | ···················· | ···················· | ···················· | ···················· |
| R | 5176 hCoV-19/England/QEUH-B0DDB6/2020\|EPI_ISL_641973\|20 | ···················· | ···················· | ···················· | ···················· | ···················· | ···················· |
| R | 5177 hCoV-19/England/QEUH-B0F8E0/2020\|EPI_ISL_641974\|20 | ···················· | ···················· | ···················· | ···················· | ···················· | ···················· |
| R | 5178 hCoV-19/England/QEUH-B0F2E2/2020\|EPI_ISL_641975\|20 | ···················· | ···················· | ···················· | ···················· | ···················· | ···················· |
| R | 5179 hCoV-19/England/QEUH-B10F8D/2020\|EPI_ISL_641976\|20 | ···················· | ···················· | ···················· | ···················· | ···················· | ···················· |
| R | 5180 hCoV-19/England/QEUH-B0F983/2020\|EPI_ISL_641977\|20 | ···················· | ···················· | ···················· | ···················· | ···················· | ···················· |
| R | 5181 hCoV-19/England/QEUH-B0DC9B/2020\|EPI_ISL_641978\|20 | ···················· | ···················· | ···················· | ···················· | ···················· | ···················· |
| R | 5182 hCoV-19/England/QEUH-B0DE68/2020\|EPI_ISL_641979\|20 | ···················· | ···················· | ···················· | ···················· | ···················· | ···················· |
| R | 5183 hCoV-19/England/QEUH-B0E438/2020\|EPI_ISL_641980\|20 | ···················· | ···················· | ···················· | ···················· | ···················· | ···················· |
| R | 5184 hCoV-19/England/QEUH-B0E9DF/2020\|EPI_ISL_641981\|20 | ···················· | ···················· | ···················· | ···················· | ······T············· | ···················· |
| R | 5185 hCoV-19/England/QEUH-B0E104/2020\|EPI_ISL_641982\|20 | ···················· | ···················· | ···················· | ···················· | ···················· | ···················· |
| R | 5186 hCoV-19/England/QEUH-B10FE7/2020\|EPI_ISL_641983\|20 | ···················· | ···················· | ···················· | ···················· | ···················· | ···················· |
| R | 5187 hCoV-19/England/QEUH-B0E9A2/2020\|EPI_ISL_641984\|20 | ···················· | ···················· | ···················· | ···················· | ······T············· | ···················· |
| R | 5188 hCoV-19/England/QEUH-B0E289/2020\|EPI_ISL_641985\|20 | ···················· | ···················· | ···················· | ···················· | ···················· | ···················· |
| R | 5189 hCoV-19/England/QEUH-B0EC12/2020\|EPI_ISL_641988\|20 | ···················· | ···················· | ···················· | ···················· | ···················· | ···················· |
| R | 5190 hCoV-19/England/QEUH-B0E5BD/2020\|EPI_ISL_641989\|20 | ···················· | ···················· | ···················· | ···················· | ···················· | ···················· |
| R | 5191 hCoV-19/England/QEUH-B0F3FE/2020\|EPI_ISL_641990\|20 | ···················· | ···················· | ···················· | ···················· | ···················· | ···················· |
| R | 5192 hCoV-19/England/QEUH-B0E34A/2020\|EPI_ISL_641991\|20 | ···················· | ···················· | ···················· | ···················· | ······T············· | ···················· |
| R | 5193 hCoV-19/England/QEUH-B0E526/2020\|EPI_ISL_641992\|20 | ···················· | ···················· | ···················· | ···················· | ···················· | ···················· |
| R | 5194 hCoV-19/England/QEUH-B0FB6F/2020\|EPI_ISL_641993\|20 | ···················· | ···················· | ···················· | ···················· | ···················· | ···················· |
| R | 5195 hCoV-19/England/QEUH-B0DED1/2020\|EPI_ISL_641994\|20 | ···················· | ···················· | ···················· | ···················· | ···················· | ···················· |
| R | 5196 hCoV-19/England/QEUH-B0E966/2020\|EPI_ISL_641996\|20 | ···················· | ···················· | ···················· | ···················· | ···················· | ···················· |
| R | 5197 hCoV-19/England/QEUH-B0E483/2020\|EPI_ISL_641997\|20 | ···················· | ·A·················· | ···················· | ···················· | ···················· | ···A················ |
| R | 5198 hCoV-19/England/QEUH-B0DE1D/2020\|EPI_ISL_641998\|20 | ···················· | ···················· | ···················· | ···················· | ···················· | ···················· |
| R | 5199 hCoV-19/England/QEUH-B0DD89/2020\|EPI_ISL_641999\|20 | ···················· | ···················· | ···················· | ···················· | ···················· | ···················· |
| R | 5200 hCoV-19/Scotland/QEUH-AAE7C9/2020\|EPI_ISL_644101\|2 | ···················· | ···················· | ···················· | ···················· | ···················· | ···················· |
| R | 5201 hCoV-19/England/ALDP-AA2A80/2020\|EPI_ISL_644103\|20 | ···················· | ···················· | ···················· | ···················· | ···················· | ···················· |
| R | 5202 hCoV-19/England/ALDP-AA0A0A/2020\|EPI_ISL_644104\|20 | ···················· | ···················· | ···················· | ···················· | ···················· | ···················· |
| R | 5203 hCoV-19/England/ALDP-AA29ED/2020\|EPI_ISL_644105\|20 | ···················· | ···················· | ···················· | ···················· | ···················· | ···················· |
| R | 5204 hCoV-19/England/ALDP-AA2A62/2020\|EPI_ISL_644106\|20 | ···················· | ···················· | ···················· | ···················· | ···················· | ···················· |
| R | 5205 hCoV-19/England/QEUH-AAF0EB/2020\|EPI_ISL_644107\|20 | ···················· | ···················· | ···················· | ···················· | ···················· | ···················· |
| R | 5206 hCoV-19/England/ALDP-AA0985/2020\|EPI_ISL_644108\|20 | ···················· | ···················· | ···················· | ···················· | ···················· | ···················· |
| R | 5207 hCoV-19/England/ALDP-AA4ABB/2020\|EPI_ISL_644109\|20 | ···················· | ···················· | ···················· | ···················· | ···················· | ···················· |
| R | 5208 hCoV-19/England/ALDP-AA4A51/2020\|EPI_ISL_644110\|20 | ···················· | ···················· | ···················· | ···················· | ···················· | ···················· |
| R | 5209 hCoV-19/England/ALDP-AA09D0/2020\|EPI_ISL_644111\|20 | ···················· | ···················· | ···················· | ···················· | ······T············· | ···················· |
| R | 5210 hCoV-19/Scotland/QEUH-AAE680/2020\|EPI_ISL_644112\|2 | ···················· | ···················· | ···················· | ···················· | ···················· | ···················· |
| R | 5211 hCoV-19/England/ALDP-AA4DA3/2020\|EPI_ISL_644113\|20 | ···················· | ···················· | ···················· | ···················· | ···················· | ···················· |
| R | 5212 hCoV-19/England/CAMC-AAFF2A/2020\|EPI_ISL_644117\|20 | ···················· | ···················· | ···················· | ···················· | ···················· | ···················· |
| R | 5213 hCoV-19/England/ALDP-AC1283/2020\|EPI_ISL_644118\|20 | ···················· | ···················· | ···················· | ···················· | ···················· | ···················· |
| R | 5214 hCoV-19/England/MILK-ABC142/2020\|EPI_ISL_644120\|20 | ···················· | ···················· | ···················· | ···················· | ···················· | ···················· |
| R | 5215 hCoV-19/England/CAMC-B374CD/2020\|EPI_ISL_645400\|20 | ···················· | ···················· | ···················· | ···················· | ···················· | ···················· |
| R | 5216 hCoV-19/England/CAMC-B36060/2020\|EPI_ISL_645401\|20 | ···················· | ···················· | ···················· | ···················· | ···················· | ···················· |
| R | 5217 hCoV-19/England/CAMC-B3725A/2020\|EPI_ISL_645402\|20 | ···················· | ···················· | ···················· | ···················· | ···················· | ···················· |
| R | 5218 hCoV-19/England/CAMC-B36051/2020\|EPI_ISL_645403\|20 | ···················· | ···················· | ···················· | ···················· | ···················· | ···················· |
| R | 5219 hCoV-19/England/CAMC-B373DF/2020\|EPI_ISL_645404\|20 | ···················· | ···················· | ···················· | ···················· | ······T············· | ···················· |
| R | 5220 hCoV-19/England/MILK-B3D254/2020\|EPI_ISL_645405\|20 | ···················· | ···················· | ···················· | ···················· | ···················· | ···················· |
| R | 5221 hCoV-19/England/MILK-B3D6B2/2020\|EPI_ISL_645406\|20 | ···················· | ···················· | ···················· | ···················· | ······T············· | ···················· |
| R | 5222 hCoV-19/England/CAMC-B35E58/2020\|EPI_ISL_645407\|20 | ···················· | ···················· | ···················· | ···················· | ···················· | ···················· |
| R | 5223 hCoV-19/England/CAMC-B375CA/2020\|EPI_ISL_645408\|20 | ···················· | ···················· | ···················· | ···················· | ···················· | ···················· |
| R | 5224 hCoV-19/England/CAMC-B36297/2020\|EPI_ISL_645409\|20 | ···················· | ···················· | ···················· | ···················· | ···················· | ···················· |
| R | 5225 hCoV-19/England/CAMC-B372D2/2020\|EPI_ISL_645412\|20 | ···················· | ···················· | ···················· | ···················· | ···················· | ···················· |
| R | 5226 hCoV-19/England/CAMC-B3615E/2020\|EPI_ISL_645413\|20 | ···················· | ···················· | ···················· | ···················· | ···················· | ···················· |
| R | 5227 hCoV-19/England/CAMC-B366AA/2020\|EPI_ISL_645414\|20 | ···················· | ···················· | ···················· | ···················· | ···················· | ···················· |
| R | 5228 hCoV-19/England/CAMC-B368A4/2020\|EPI_ISL_645427\|20 | ···················· | ···················· | ···················· | ···················· | ···················· | ···················· |
| R | 5229 hCoV-19/England/CAMC-B36886/2020\|EPI_ISL_645428\|20 | ···················· | ···················· | ···················· | ···················· | ···················· | ···················· |
| R | 5230 hCoV-19/England/CAMC-B36482/2020\|EPI_ISL_645429\|20 | ···················· | ·A·················· | ···················· | ···················· | ···················· | ···A················ |
| R | 5231 hCoV-19/England/CAMC-B361E5/2020\|EPI_ISL_645430\|20 | ···················· | ···················· | ···················· | ···················· | ···················· | ···················· |
| R | 5232 hCoV-19/England/CAMC-B36789/2020\|EPI_ISL_645431\|20 | ···················· | ···················· | ···················· | ···················· | ···················· | ···················· |
| R | 5233 hCoV-19/England/CAMC-B362F1/2020\|EPI_ISL_645432\|20 | ···················· | ···················· | ···················· | ···················· | ···················· | ···················· |
| R | 5234 hCoV-19/England/CAMC-B37542/2020\|EPI_ISL_645433\|20 | ···················· | ···················· | ···················· | ···················· | ···················· | ···················· |
| R | 5235 hCoV-19/England/CAMC-B37409/2020\|EPI_ISL_645434\|20 | ···················· | ···················· | ···················· | ···················· | ···················· | ···················· |
| R | 5236 hCoV-19/England/CAMC-B36965/2020\|EPI_ISL_645435\|20 | ···················· | ···················· | ···················· | ···················· | ···················· | ···················· |
| R | 5237 hCoV-19/England/CAMC-B3630D/2020\|EPI_ISL_645437\|20 | ···················· | ···················· | ···················· | ···················· | ···················· | ···················· |
| R | 5238 hCoV-19/England/CAMC-B367E3/2020\|EPI_ISL_645439\|20 | ···················· | ···················· | ···················· | ···················· | ···················· | ···················· |
| R | 5239 hCoV-19/England/MILK-B3D1C0/2020\|EPI_ISL_645441\|20 | ···················· | ···················· | ···················· | ···················· | ···················· | ···················· |
| R | 5240 hCoV-19/England/CAMC-B371F3/2020\|EPI_ISL_645442\|20 | ···················· | ···················· | ···················· | ···················· | ···················· | ···················· |
| R | 5241 hCoV-19/England/CAMC-B36385/2020\|EPI_ISL_645443\|20 | ···················· | ···················· | ···················· | ···················· | ···················· | ···················· |
| R | 5242 hCoV-19/England/CAMC-B36A35/2020\|EPI_ISL_645444\|20 | ···················· | ···················· | ···················· | ···················· | ···················· | ···················· |
| R | 5243 hCoV-19/England/CAMC-B36367/2020\|EPI_ISL_645445\|20 | ···················· | ···················· | ···················· | ···················· | ···················· | ···················· |
| R | 5244 hCoV-19/England/CAMC-B36895/2020\|EPI_ISL_645446\|20 | ···················· | ···················· | ···················· | ···················· | ···················· | ···················· |
| R | 5245 hCoV-19/England/CAMC-B3732A/2020\|EPI_ISL_645448\|20 | ···················· | ···················· | ···················· | ···················· | ···················· | ···················· |
| R | 5246 hCoV-19/England/CAMC-B37533/2020\|EPI_ISL_645449\|20 | ···················· | ···················· | ···················· | ···················· | ···················· | ···················· |
| R | 5247 hCoV-19/England/CAMC-B36200/2020\|EPI_ISL_645450\|20 | ···················· | ···················· | ···················· | ···················· | ···················· | ···················· |
| R | 5248 hCoV-19/England/CAMC-B3619A/2020\|EPI_ISL_645451\|20 | ···················· | ···················· | ···················· | ···················· | ···················· | ···················· |
| R | 5249 hCoV-19/England/ALDP-B3EDE9/2020\|EPI_ISL_645452\|20 | ···················· | ···················· | ···················· | ···················· | ···················· | ···················· |
| R | 5250 hCoV-19/England/MILK-B3D51F/2020\|EPI_ISL_645453\|20 | ···················· | ···················· | ···················· | ···················· | ···················· | ···················· |
| R | 5251 hCoV-19/England/CAMC-B3716C/2020\|EPI_ISL_645454\|20 | ···················· | ···················· | ···················· | ···················· | ···················· | ···················· |

R 5252 hCoV-19/England/CAMC-B3722D/2020|EPI_ISL_645455|20 ···················· ···················· ····················· ···················· ···················· ····················
R 5253 hCoV-19/England/CAMC-B3682C/2020|EPI_ISL_645457|20 ···················· ···················· ····················· ···················· ······T············· ····················
R 5254 hCoV-19/England/CAMC-B360BB/2020|EPI_ISL_645458|20 ···················· ···················· ····················· ···················· ······T············· ····················
R 5255 hCoV-19/England/ALDP-B3EB0D/2020|EPI_ISL_645459|20 ···················· ···················· ····················· ···················· ···················· ····················
R 5256 hCoV-19/England/CAMC-B374EB/2020|EPI_ISL_645460|20 ···················· ···················· ····················· ···················· ···················· ····················
R 5257 hCoV-19/England/CAMC-B36604/2020|EPI_ISL_645461|20 ···················· ·A·················· ····················· ···················· ···················· ···A················
R 5258 hCoV-19/England/CAMC-B368B3/2020|EPI_ISL_645462|20 ···················· ···················· ····················· ···················· ···················· ····················
R 5259 hCoV-19/England/CAMC-B361C7/2020|EPI_ISL_645463|20 ···················· ···················· ····················· ···················· ···················· ····················
R 5260 hCoV-19/England/ALDP-B3ED52/2020|EPI_ISL_645464|20 ···················· ···················· ····················· ···················· ···················· ····················
R 5261 hCoV-19/England/CAMC-B37515/2020|EPI_ISL_645465|20 ···················· ···················· ····················· ···················· ···················· ····················
R 5262 hCoV-19/England/CAMC-B37111/2020|EPI_ISL_645466|20 ···················· ···················· ····················· ···················· ···················· ····················
R 5263 hCoV-19/England/CAMC-B3759D/2020|EPI_ISL_645467|20 ···················· ···················· ····················· ···················· ······T············· ····················
R 5264 hCoV-19/England/CAMC-B370D8/2020|EPI_ISL_645468|20 ···················· ···················· ····················· ···················· ···················· ····················
R 5265 hCoV-19/England/CAMC-B36419/2020|EPI_ISL_645469|20 ···················· ···················· ····················· ···················· ···················· ····················
R 5266 hCoV-19/England/CAMC-B375BB/2020|EPI_ISL_645470|20 ···················· ···················· ····················· ···················· ···················· ····················
R 5267 hCoV-19/England/CAMC-B37102/2020|EPI_ISL_645472|20 ···················· ···················· ····················· ···················· ···················· ····················
R 5268 hCoV-19/England/ALDP-B3E657/2020|EPI_ISL_645473|20 ···················· ···················· ····················· ···················· ···················· ····················
R 5269 hCoV-19/England/CAMC-B3680E/2020|EPI_ISL_645474|20 ···················· ···················· ····················· ···················· ···················· ····················
R 5270 hCoV-19/England/CAMC-B35F37/2020|EPI_ISL_645475|20 ···················· ···················· ····················· ···················· ···················· ····················
R 5271 hCoV-19/England/CAMC-B36279/2020|EPI_ISL_645476|20 ···················· ···················· ····················· ···················· ···················· ····················
R 5272 hCoV-19/England/MILK-B3D4D6/2020|EPI_ISL_645477|20 ···················· ···················· ····················· ···················· ···················· ····················
R 5273 hCoV-19/England/CAMC-B361B8/2020|EPI_ISL_645478|20 ···················· ·A·················· ····················· ···················· ···················· ····················
R 5274 hCoV-19/England/CAMC-B365DA/2020|EPI_ISL_645479|20 ···················· ···················· ····················· ···················· ···················· ····················
R 5275 hCoV-19/England/CAMC-B3709C/2020|EPI_ISL_645480|20 ···················· ···················· ····················· ···················· ···················· ····················
R 5276 hCoV-19/England/CAMC-B3691A/2020|EPI_ISL_645482|20 ···················· ···················· ····················· ···················· ···················· ····················
R 5277 hCoV-19/England/CAMC-B368FF/2020|EPI_ISL_645483|20 ···················· ···················· ····················· ···················· ···················· ···A················
R 5278 hCoV-19/England/CAMC-B37445/2020|EPI_ISL_645484|20 ···················· ···················· ····················· ···················· ···················· ····················
R 5279 hCoV-19/England/CAMC-B3621F/2020|EPI_ISL_645485|20 ···················· ···················· ····················· ···················· ···················· ····················
R 5280 hCoV-19/England/CAMC-B37436/2020|EPI_ISL_645486|20 ···················· ···················· ····················· ···················· ···················· ····················
R 5281 hCoV-19/England/CAMC-B36437/2020|EPI_ISL_645487|20 ···················· ···················· ····················· ···················· ···················· ····················
R 5282 hCoV-19/England/CAMC-B36103/2020|EPI_ISL_645488|20 ···················· ···················· ····················· ···················· ···················· ····················
R 5283 hCoV-19/England/MILK-B3D694/2020|EPI_ISL_645489|20 ···················· ·A·················· ····················· ···················· ···················· ···A················
R 5284 hCoV-19/England/QEUH-B2B927/2020|EPI_ISL_646700|20 ···················· ···················· ····················· ···················· ···················· ····················
R 5285 hCoV-19/England/QEUH-B2B6B7/2020|EPI_ISL_646701|20 ···················· ···················· ····················· ···················· ···················· ····················
R 5286 hCoV-19/England/ALDP-B26458/2020|EPI_ISL_646702|20 ···················· ···················· ····················· ···················· ···················· ····················
R 5287 hCoV-19/England/QEUH-B2B3ED/2020|EPI_ISL_646703|20 ···················· ···················· ····················· ···················· ···················· ····················
R 5288 hCoV-19/England/QEUH-B2CAD8/2020|EPI_ISL_646704|20 ···················· ···················· ····················· ···················· ···················· ····················
R 5289 hCoV-19/England/ALDP-B2635B/2020|EPI_ISL_646706|20 ···················· ···················· ····················· ···················· ···················· ····················
R 5290 hCoV-19/England/ALDP-B25EE2/2020|EPI_ISL_646707|20 ···················· ···················· ····················· ···················· ···················· ····················
R 5291 hCoV-19/England/ALDP-B25240/2020|EPI_ISL_646708|20 ···················· ···················· ····················· ···················· ···················· ····················
R 5292 hCoV-19/England/ALDP-B24E89/2020|EPI_ISL_646710|20 ···················· ···················· ····················· ···················· ···················· ····················
R 5293 hCoV-19/England/ALDP-B25ED3/2020|EPI_ISL_646711|20 ···················· ···················· ····················· ···················· ···················· ····················
R 5294 hCoV-19/England/ALDP-B25170/2020|EPI_ISL_646712|20 ···················· ·A·················· ····················· ···················· ···················· ····················
R 5295 hCoV-19/England/ALDP-B26133/2020|EPI_ISL_646713|20 ···················· ···················· ····················· ···················· ···················· ····················
R 5296 hCoV-19/England/QEUH-B2B2E0/2020|EPI_ISL_646714|20 ···················· ···················· ····················· ···················· ···················· ····················
R 5297 hCoV-19/England/ALDP-B26027/2020|EPI_ISL_646715|20 ···················· ···················· ····················· ···················· ···················· ····················
R 5298 hCoV-19/England/ALDP-B24E5C/2020|EPI_ISL_646716|20 ···················· ·A·················· ····················· ···················· ···················· ····················
R 5299 hCoV-19/England/QEUH-B2B480/2020|EPI_ISL_646717|20 ···················· ···················· ····················· ···················· ···················· ····················
R 5300 hCoV-19/England/ALDP-B24DF5/2020|EPI_ISL_646718|20 ···················· ···················· ····················· ···················· ···················· ····················
R 5301 hCoV-19/England/ALDP-B251AD/2020|EPI_ISL_646719|20 ···················· ···················· ····················· ···················· ···················· ····················
R 5302 hCoV-19/England/QEUH-B2B374/2020|EPI_ISL_646720|20 ···················· ···················· ····················· ···················· ···················· ····················
R 5303 hCoV-19/England/ALDP-B24E10/2020|EPI_ISL_646721|20 ···················· ···················· ····················· ···················· ···················· ····················
R 5304 hCoV-19/England/QEUH-B2C9F9/2020|EPI_ISL_646723|20 ···················· ·A·················· ····················· ···················· ···················· ···A················
R 5305 hCoV-19/England/ALDP-B2631F/2020|EPI_ISL_646724|20 ···················· ···················· ····················· ···················· ···················· ····················
R 5306 hCoV-19/England/QEUH-B2B3B0/2020|EPI_ISL_646725|20 ···················· ···················· ····················· ···················· ···················· ····················
R 5307 hCoV-19/England/QEUH-B2C8B0/2020|EPI_ISL_646726|20 ···················· ···················· ····················· ···················· ···················· ····················
R 5308 hCoV-19/England/ALDP-B26449/2020|EPI_ISL_646727|20 ···················· ···················· ····················· ···················· ···················· ····················
R 5309 hCoV-19/England/QEUH-B2B2C2/2020|EPI_ISL_646728|20 ···················· ···················· ····················· ···················· ···················· ····················
R 5310 hCoV-19/England/QEUH-B2B49F/2020|EPI_ISL_646729|20 ···················· ···················· ····················· ···················· ···················· ····················
R 5311 hCoV-19/England/QEUH-B2B82A/2020|EPI_ISL_646730|20 ···················· ···················· ····················· ···················· ···················· ····················
R 5312 hCoV-19/England/ALDP-B24EB6/2020|EPI_ISL_646731|20 ···················· ·A·················· ····················· ···················· ···················· ····················
R 5313 hCoV-19/England/QEUH-B2CA32/2020|EPI_ISL_646732|20 ···················· ···················· ····················· ···················· ···················· ····················
R 5314 hCoV-19/England/QEUH-B2B602/2020|EPI_ISL_646733|20 ···················· ···················· ····················· ···················· ···················· ····················
R 5315 hCoV-19/England/ALDP-B2641C/2020|EPI_ISL_646734|20 ···················· ···················· ····················· ···················· ···················· ····················
R 5316 hCoV-19/England/QEUH-B2B9FA/2020|EPI_ISL_646735|20 ···················· ···················· ····················· ···················· ···················· ····················
R 5317 hCoV-19/England/ALDP-B25F2B/2020|EPI_ISL_646736|20 ···················· ···················· ····················· ···················· ···················· ····················
R 5318 hCoV-19/England/ALDP-B2636A/2020|EPI_ISL_646737|20 ···················· ···················· ····················· ···················· ···················· ····················
R 5319 hCoV-19/Wales/ALDP-B24F68/2020|EPI_ISL_646738|2020 ···················· ···················· ····················· ···················· ···················· ····················
R 5320 hCoV-19/England/ALDP-B2529B/2020|EPI_ISL_646739|20 ···················· ···················· ····················· ···················· ···················· ····················
R 5321 hCoV-19/England/ALDP-B25F76/2020|EPI_ISL_646743|20 ···················· ···················· ····················· ···················· ···················· ····················
R 5322 hCoV-19/England/QEUH-B2BA51/2020|EPI_ISL_646745|20 ···················· ···················· ····················· ···················· ···················· ····················
R 5323 hCoV-19/England/ALDP-B24D40/2020|EPI_ISL_646746|20 ···················· ···················· ····················· ···················· ···················· ····················
R 5324 hCoV-19/England/ALDP-B25FFE/2020|EPI_ISL_646758|20 ···················· ···················· ····················· ···················· ···················· ····················
R 5325 hCoV-19/England/QEUH-B2C883/2020|EPI_ISL_646759|20 ···················· ···················· ····················· ···················· ···················· ····················
R 5326 hCoV-19/England/ALDP-B251F8/2020|EPI_ISL_646760|20 ···················· ···················· ····················· ···················· ···················· ····················
R 5327 hCoV-19/England/ALDP-B260DC/2020|EPI_ISL_646761|20 ···················· ···················· ····················· ···················· ···················· ····················
R 5328 hCoV-19/England/ALDP-B25019/2020|EPI_ISL_646762|20 ···················· ···················· ····················· ···················· ···················· ····················
R 5329 hCoV-19/England/ALDP-B25F94/2020|EPI_ISL_646763|20 ···················· ···················· ····················· ···················· ···················· ····················
R 5330 hCoV-19/England/QEUH-B2C874/2020|EPI_ISL_646764|20 ···················· ···················· ····················· ···················· ···················· ····················
R 5331 hCoV-19/England/QEUH-B2C9BD/2020|EPI_ISL_646765|20 ···················· ···················· ····················· ···················· ···················· ····················

| | | | | | | | |
|---|---|---|---|---|---|---|---|
| R | 5332 hCoV-19/England/QEUH-B2B3A1/2020\|EPI_ISL_646766\|20 | ···················· | ···················· | ···················· | ···················· | ···················· | ···················· |
| R | 5333 hCoV-19/England/ALDP-B24F0E/2020\|EPI_ISL_646767\|20 | ···················· | ···················· | ···················· | ···················· | ···················· | ···················· |
| R | 5334 hCoV-19/England/ALDP-B263C4/2020\|EPI_ISL_646768\|20 | ···················· | ···················· | ···················· | ···················· | ···················· | ···················· |
| R | 5335 hCoV-19/England/ALDP-B25161/2020\|EPI_ISL_646771\|20 | ···················· | ·A·················· | ···················· | ···················· | ···················· | ···················· |
| R | 5336 hCoV-19/England/QEUH-B2BACA/2020\|EPI_ISL_646772\|20 | ···················· | ···················· | ···················· | ···················· | ···················· | ···················· |
| R | 5337 hCoV-19/England/ALDP-B2640D/2020\|EPI_ISL_646773\|20 | ···················· | ···················· | ···················· | ···················· | ···················· | ···················· |
| R | 5338 hCoV-19/England/QEUH-B2B505/2020\|EPI_ISL_646774\|20 | ···················· | ···················· | ···················· | ···················· | ···················· | ···················· |
| R | 5339 hCoV-19/England/QEUH-B2C962/2020\|EPI_ISL_646775\|20 | ···················· | ···················· | ···················· | ···················· | ···················· | ···················· |
| R | 5340 hCoV-19/England/ALDP-B24E4D/2020\|EPI_ISL_646776\|20 | ···················· | ···················· | ···················· | ···················· | ···················· | ···················· |
| R | 5341 hCoV-19/England/QEUH-B2B7B4/2020\|EPI_ISL_646777\|20 | ···················· | ···················· | ···················· | ···················· | ···················· | ···················· |
| R | 5342 hCoV-19/England/QEUH-B2B31A/2020\|EPI_ISL_646778\|20 | ···················· | ···················· | ···················· | ···················· | ···················· | ···················· |
| R | 5343 hCoV-19/England/QEUH-B2BAAC/2020\|EPI_ISL_646779\|20 | ···················· | ···················· | ···················· | ···················· | ···················· | ···················· |
| R | 5344 hCoV-19/England/QEUH-B2B5BA/2020\|EPI_ISL_646781\|20 | ···················· | ···················· | ···················· | ···················· | ···················· | ···················· |
| R | 5345 hCoV-19/England/ALDP-B25222/2020\|EPI_ISL_646782\|20 | ···················· | ···················· | ···················· | ···················· | ···················· | ···················· |
| R | 5346 hCoV-19/England/QEUH-B2C935/2020\|EPI_ISL_646783\|20 | ···················· | ···················· | ···················· | ···················· | ···················· | ···················· |
| R | 5347 hCoV-19/England/QEUH-B2B73C/2020\|EPI_ISL_646785\|20 | ···················· | ···················· | ···················· | ···················· | ···················· | ···················· |
| R | 5348 hCoV-19/England/QEUH-B2B2D1/2020\|EPI_ISL_646786\|20 | ···················· | ···················· | ···················· | ···················· | ···················· | ···················· |
| R | 5349 hCoV-19/England/QEUH-B2B7E1/2020\|EPI_ISL_646788\|20 | ···················· | ···················· | ···················· | ···················· | ···················· | ···················· |
| R | 5350 hCoV-19/England/ALDP-B23A68/2020\|EPI_ISL_646789\|20 | ···················· | ···················· | ···················· | ···················· | ···················· | ···················· |
| R | 5351 hCoV-19/England/ALDP-B24812/2020\|EPI_ISL_646790\|20 | ···················· | ···················· | ···················· | ···················· | ···················· | ···················· |
| R | 5352 hCoV-19/England/QEUH-B2D72B/2020\|EPI_ISL_646792\|20 | ···················· | ···················· | ···················· | ···················· | ···················· | ···················· |
| R | 5353 hCoV-19/England/ALDP-B24B82/2020\|EPI_ISL_646793\|20 | ···················· | ···················· | ···················· | ···················· | ···················· | ···················· |
| R | 5354 hCoV-19/England/QEUH-B2ED37/2020\|EPI_ISL_646794\|20 | ···················· | ···················· | ···················· | ···················· | ···················· | ···················· |
| R | 5355 hCoV-19/England/ALDP-B2435D/2020\|EPI_ISL_646795\|20 | ···················· | ···················· | ···················· | ···················· | ···················· | ···················· |
| R | 5356 hCoV-19/England/ALDP-B2396B/2020\|EPI_ISL_646796\|20 | ···················· | ···················· | ···················· | ···················· | ···················· | ···················· |
| R | 5357 hCoV-19/England/ALDP-B2489A/2020\|EPI_ISL_646797\|20 | ···················· | ···················· | ···················· | ···················· | ···················· | ···················· |
| R | 5358 hCoV-19/England/QEUH-B2DA8C/2020\|EPI_ISL_646798\|20 | ···················· | ···················· | ···················· | ···················· | ···················· | ···················· |
| R | 5359 hCoV-19/England/ALDP-B23AB3/2020\|EPI_ISL_646799\|20 | ···················· | ···················· | ···················· | ···················· | ···················· | ···················· |
| R | 5360 hCoV-19/USA/CA-ALSR-4388/2020\|EPI_ISL_649001\|2020- | ···················· | ·A·················· | ···················· | ···················· | ···················· | ···················· |
| R | 5361 hCoV-19/USA/CA-ALSR-4391/2020\|EPI_ISL_649004\|2020- | ···················· | ·A·················· | ···················· | ···················· | ···················· | ···················· |
| R | 5362 hCoV-19/USA/CA-ALSR-4395/2020\|EPI_ISL_649005\|2020- | ···················· | ·A·················· | ···················· | ···················· | ···················· | ···················· |
| R | 5363 hCoV-19/USA/CA-ALSR-4398/2020\|EPI_ISL_649006\|2020- | ···················· | ···················· | ···················· | ···················· | ···················· | ···················· |
| R | 5364 hCoV-19/USA/CA-ALSR-4399/2020\|EPI_ISL_649007\|2020- | ···················· | ·A·················· | ···················· | ···················· | ···················· | ···················· |
| R | 5365 hCoV-19/USA/CA-ALSR-4400/2020\|EPI_ISL_649008\|2020- | ···················· | ·A·················· | ···················· | ···················· | ···················· | ···················· |
| R | 5366 hCoV-19/USA/CA-ALSR-4401/2020\|EPI_ISL_649009\|2020- | ···················· | ·A·················· | ···················· | ···················· | ···················· | ···················· |
| R | 5367 hCoV-19/USA/CA-ALSR-4413/2020\|EPI_ISL_649016\|2020- | ···················· | ···················· | ···················· | ···················· | ···················· | ···················· |
| R | 5368 hCoV-19/USA/CA-ALSR-4417/2020\|EPI_ISL_649017\|2020- | ···················· | ·A·················· | ···················· | ···················· | ···················· | ···················· |
| R | 5369 hCoV-19/USA/CA-ALSR-4419/2020\|EPI_ISL_649018\|2020- | ···················· | ·A·················· | ···················· | ···················· | ···················· | ···················· |
| R | 5370 hCoV-19/USA/CA-ALSR-4425/2020\|EPI_ISL_649020\|2020- | ···················· | ·A·················· | ···················· | ···················· | ···················· | ···················· |
| R | 5371 hCoV-19/USA/CA-ALSR-4428/2020\|EPI_ISL_649022\|2020- | ···················· | ·A·················· | ···················· | ···················· | ···················· | ···················· |
| R | 5372 hCoV-19/USA/CA-ALSR-4429/2020\|EPI_ISL_649023\|2020- | ···················· | ·A·················· | ···················· | ···················· | ···················· | ···················· |
| R | 5373 hCoV-19/USA/CA-ALSR-4431/2020\|EPI_ISL_649025\|2020- | ···················· | ·A·················· | ···················· | ···················· | ···················· | ···················· |
| R | 5374 hCoV-19/USA/CA-ALSR-4433/2020\|EPI_ISL_649027\|2020- | ···················· | ·A·················· | ···················· | ···················· | ···················· | ···················· |
| R | 5375 hCoV-19/USA/CA-ALSR-4435/2020\|EPI_ISL_649028\|2020- | ···················· | ·A·················· | ···················· | ···················· | ···················· | ···················· |
| R | 5376 hCoV-19/USA/CA-ALSR-4436/2020\|EPI_ISL_649029\|2020- | ···················· | ···················· | ···················· | ···················· | ···················· | ···················· |
| R | 5378 hCoV-19/Australia/VIC12895/2020\|EPI_ISL_640501\|202 | ···················· | ···················· | ···················· | ···················· | ···················· | ···················· |
| R | 5379 hCoV-19/Australia/VIC11593/2020\|EPI_ISL_640502\|202 | ···················· | ···················· | ···················· | ···················· | ···················· | ···················· |
| R | 5380 hCoV-19/Australia/VIC13281/2020\|EPI_ISL_640503\|202 | ···················· | ···················· | ···················· | ···················· | ···················· | ···················· |
| R | 5381 hCoV-19/Australia/VIC16005/2020\|EPI_ISL_640504\|202 | ···················· | ···················· | ···················· | ···················· | ···················· | ···················· |
| R | 5382 hCoV-19/Australia/VIC12454/2020\|EPI_ISL_640505\|202 | ···················· | ···················· | ···················· | ···················· | ···················· | ···················· |
| R | 5383 hCoV-19/Australia/VIC16543/2020\|EPI_ISL_640506\|202 | ···················· | ···················· | ···················· | ···················· | ···················· | ···················· |
| R | 5384 hCoV-19/Australia/VIC12746/2020\|EPI_ISL_640507\|202 | ···················· | ···················· | ···················· | ···················· | ···················· | ···················· |
| R | 5385 hCoV-19/Australia/VIC12872/2020\|EPI_ISL_640508\|202 | ···················· | ···················· | ···················· | ···················· | ···················· | ···················· |
| R | 5386 hCoV-19/Australia/VIC16161/2020\|EPI_ISL_640510\|202 | ···················· | ···················· | ···················· | ···················· | ···················· | ···················· |
| R | 5387 hCoV-19/Australia/VIC12718/2020\|EPI_ISL_640511\|202 | ···················· | ···················· | ···················· | ···················· | ···················· | ···················· |
| R | 5388 hCoV-19/Australia/VIC12009/2020\|EPI_ISL_640513\|202 | ···················· | ···················· | ···················· | ···················· | ···················· | ···················· |
| R | 5389 hCoV-19/Australia/VIC12078/2020\|EPI_ISL_640514\|202 | ···················· | ···················· | ···················· | ···················· | ···················· | ···················· |
| R | 5391 hCoV-19/Australia/VIC16626/2020\|EPI_ISL_640517\|202 | ···················· | ···················· | ···················· | ···················· | ···················· | ···················· |
| R | 5392 hCoV-19/Australia/VIC16393/2020\|EPI_ISL_640518\|202 | ···················· | ···················· | ···················· | ···················· | ···················· | ···················· |
| R | 5393 hCoV-19/Australia/VIC16197/2020\|EPI_ISL_640519\|202 | ···················· | ···················· | ···················· | ···················· | ···················· | ···················· |
| R | 5394 hCoV-19/Australia/VIC16109/2020\|EPI_ISL_640520\|202 | ···················· | ···················· | ···················· | ···················· | ···················· | ···················· |
| R | 5396 hCoV-19/Australia/VIC16254/2020\|EPI_ISL_640523\|202 | ···················· | ···················· | ···················· | ···················· | ···················· | ···················· |
| R | 5397 hCoV-19/Australia/VIC12026/2020\|EPI_ISL_640524\|202 | ···················· | ···················· | ···················· | ···················· | ···················· | ···················· |
| R | 5399 hCoV-19/Australia/VIC12013/2020\|EPI_ISL_640529\|202 | ···················· | ···················· | ···················· | ···················· | ···················· | ···················· |
| R | 5400 hCoV-19/Australia/VIC16514/2020\|EPI_ISL_640533\|202 | ········T··········· | ···················· | ···················· | ···················· | ···················· | ···················· |
| R | 5401 hCoV-19/Australia/VIC15968/2020\|EPI_ISL_640535\|202 | ···················· | ···················· | ···················· | ···················· | ···················· | ···················· |
| R | 5402 hCoV-19/Australia/VIC12448/2020\|EPI_ISL_640536\|202 | ···················· | ···················· | ···················· | ···················· | ···················· | ···················· |
| R | 5403 hCoV-19/Australia/VIC16069/2020\|EPI_ISL_640541\|202 | ···················· | ···················· | ···················· | ···················· | ···················· | ···················· |
| R | 5404 hCoV-19/Australia/VIC12005/2020\|EPI_ISL_640544\|202 | ···················· | ···················· | ···················· | ···················· | ···················· | ···················· |
| R | 5405 hCoV-19/Australia/VIC15909/2020\|EPI_ISL_640545\|202 | ···················· | ···················· | ···················· | ···················· | ···················· | ···················· |
| R | 5406 hCoV-19/Australia/VIC12631/2020\|EPI_ISL_640546\|202 | ···················· | ···················· | ···················· | ···················· | ···················· | ···················· |
| R | 5407 hCoV-19/Australia/VIC12841/2020\|EPI_ISL_640547\|202 | ···················· | ···················· | ···················· | ···················· | ···················· | ···················· |
| R | 5408 hCoV-19/Australia/VIC16057/2020\|EPI_ISL_640548\|202 | ···················· | ···················· | ···················· | ···················· | ···················· | ···················· |
| R | 5409 hCoV-19/Australia/VIC12153/2020\|EPI_ISL_640549\|202 | ···················· | ···················· | ···················· | ···················· | ···················· | ···················· |
| R | 5410 hCoV-19/Australia/VIC12048/2020\|EPI_ISL_640550\|202 | ···················· | ···················· | ···················· | ···················· | ···················· | ···················· |
| R | 5411 hCoV-19/Australia/VIC16346/2020\|EPI_ISL_640552\|202 | ···················· | ···················· | ···················· | ···················· | ···················· | ···················· |
| R | 5412 hCoV-19/Australia/VIC13220/2020\|EPI_ISL_640553\|202 | ···················· | ···················· | ···················· | ···················· | ···················· | ···················· |
| R | 5413 hCoV-19/Australia/VIC16421/2020\|EPI_ISL_640554\|202 | ···················· | ···················· | ···················· | ···················· | ···················· | ···················· |
| R | 5414 hCoV-19/Australia/VIC12618/2020\|EPI_ISL_640555\|202 | ···················· | ···················· | ···················· | ···················· | ···················· | ···················· |
| R | 5416 hCoV-19/Australia/VIC12426/2020\|EPI_ISL_640558\|202 | ···················· | ···················· | ···················· | ···················· | ···················· | ···················· |

| | | | | | | | |
|---|---|---|---|---|---|---|---|
| R | 5418 hCoV-19/Australia/VIC12849/2020\|EPI_ISL_640560\|202 | ···················· | ···················· | ····················· | ···················· | ···················· | ···················· |
| R | 5419 hCoV-19/Australia/VIC16241/2020\|EPI_ISL_640561\|202 | ···················· | ···················· | ····················· | ···················· | ···················· | ···················· |
| R | 5421 hCoV-19/Australia/VIC16019/2020\|EPI_ISL_640566\|202 | ···················· | ···················· | ····················· | ···················· | ···················· | ···················· |
| R | 5422 hCoV-19/Australia/VIC12497/2020\|EPI_ISL_640567\|202 | ···················· | ···················· | ····················· | ···················· | ···················· | ···················· |
| R | 5423 hCoV-19/Australia/VIC12667/2020\|EPI_ISL_640569\|202 | ···················· | ···················· | ····················· | ···················· | ···················· | ···················· |
| R | 5424 hCoV-19/Australia/VIC12343/2020\|EPI_ISL_640570\|202 | ···················· | ···················· | ····················· | ···················· | ···················· | ···················· |
| R | 5426 hCoV-19/Australia/VIC12285/2020\|EPI_ISL_640572\|202 | ···················· | ···················· | ····················· | ···················· | ···················· | ···················· |
| R | 5427 hCoV-19/Australia/VIC16002/2020\|EPI_ISL_640573\|202 | ···················· | ···················· | ····················· | ···················· | ···················· | ···················· |
| R | 5428 hCoV-19/Australia/VIC12663/2020\|EPI_ISL_640574\|202 | ···················· | ···················· | ····················· | ···················· | ···················· | ···················· |
| R | 5429 hCoV-19/Australia/VIC15921/2020\|EPI_ISL_640575\|202 | ···················· | ···················· | ····················· | ···················· | ···················· | ···················· |
| R | 5430 hCoV-19/Australia/VIC12363/2020\|EPI_ISL_640576\|202 | ···················· | ···················· | ····················· | ···················· | ···················· | ···················· |
| R | 5431 hCoV-19/Australia/VIC12442/2020\|EPI_ISL_640577\|202 | ···················· | ···················· | ····················· | ···················· | ···················· | ···················· |
| R | 5433 hCoV-19/Australia/VIC12000/2020\|EPI_ISL_640579\|202 | ···················· | ···················· | ····················· | ···················· | ···················· | ···················· |
| R | 5435 hCoV-19/Australia/VIC16552/2020\|EPI_ISL_640581\|202 | ···················· | ···················· | ····················· | ···················· | ···················· | ···················· |
| R | 5436 hCoV-19/Australia/VIC13197/2020\|EPI_ISL_640582\|202 | ···················· | ···················· | ····················· | ···················· | ···················· | ···················· |
| R | 5438 hCoV-19/Australia/VIC13193/2020\|EPI_ISL_640584\|202 | ···················· | ···················· | ····················· | ···················· | ···················· | ···················· |
| R | 5440 hCoV-19/Australia/VIC16078/2020\|EPI_ISL_640587\|202 | ···················· | ···················· | ····················· | ···················· | ···················· | ···················· |
| R | 5441 hCoV-19/Australia/VIC16199/2020\|EPI_ISL_640588\|202 | ···················· | ···················· | ····················· | ···················· | ···················· | ···················· |
| R | 5442 hCoV-19/Australia/VIC16040/2020\|EPI_ISL_640589\|202 | ···················· | ···················· | ····················· | ···················· | ···················· | ···················· |
| R | 5443 hCoV-19/Australia/VIC16384/2020\|EPI_ISL_640591\|202 | ···················· | ···················· | ····················· | ···················· | ···················· | ···················· |
| R | 5444 hCoV-19/Australia/VIC12259/2020\|EPI_ISL_640592\|202 | ···················· | ···················· | ····················· | ···················· | ···················· | ···················· |
| R | 5446 hCoV-19/Australia/VIC12799/2020\|EPI_ISL_640594\|202 | ···················· | ···················· | ····················· | ···················· | ···················· | ···················· |
| R | 5447 hCoV-19/Australia/VIC12095/2020\|EPI_ISL_640595\|202 | ···················· | ···················· | ····················· | ···················· | ···················· | ···················· |
| R | 5448 hCoV-19/Australia/VIC12411/2020\|EPI_ISL_640596\|202 | ···················· | ···················· | ····················· | ···················· | ···················· | ···················· |
| R | 5449 hCoV-19/Australia/VIC12777/2020\|EPI_ISL_640597\|202 | ···················· | ···················· | ····················· | ···················· | ···················· | ···················· |
| R | 5450 hCoV-19/England/QEUH-B10293/2020\|EPI_ISL_641800\|20 | ···················· | ···················· | ····················· | ···················· | ···················· | ···················· |
| R | 5451 hCoV-19/England/QEUH-B12F6D/2020\|EPI_ISL_641802\|20 | ···················· | ···················· | ····················· | ···················· | ···················· | ···················· |
| R | 5452 hCoV-19/England/QEUH-B10794/2020\|EPI_ISL_641803\|20 | ···················· | ···················· | ····················· | ···················· | ···················· | ···················· |
| R | 5453 hCoV-19/England/QEUH-B13FE4/2020\|EPI_ISL_641804\|20 | ···················· | ···················· | ····················· | ···················· | ···················· | ···················· |
| R | 5454 hCoV-19/England/QEUH-B13430/2020\|EPI_ISL_641805\|20 | ···················· | ···················· | ····················· | ···················· | ···················· | ···················· |
| R | 5455 hCoV-19/England/QEUH-B108BF/2020\|EPI_ISL_641806\|20 | ···················· | ···················· | ····················· | ···················· | ···················· | ···················· |
| R | 5456 hCoV-19/England/QEUH-B12D36/2020\|EPI_ISL_641807\|20 | ···················· | ···················· | ····················· | ···················· | ···················· | ···················· |
| R | 5457 hCoV-19/England/QEUH-B133D9/2020\|EPI_ISL_641808\|20 | ···················· | ···················· | ····················· | ···················· | ···················· | ···················· |
| R | 5458 hCoV-19/England/QEUH-B145B4/2020\|EPI_ISL_641809\|20 | ···················· | ···················· | ····················· | ···················· | ···················· | ···················· |
| R | 5459 hCoV-19/England/QEUH-B13BD1/2020\|EPI_ISL_641810\|20 | ···················· | ·A·················· | ····················· | ···················· | ···················· | ···················· |
| R | 5460 hCoV-19/England/QEUH-B13AD4/2020\|EPI_ISL_641811\|20 | ···················· | ···················· | ····················· | ···················· | ···················· | ···················· |
| R | 5461 hCoV-19/England/QEUH-B12F30/2020\|EPI_ISL_641812\|20 | ···················· | ···················· | ····················· | ···················· | ···················· | ···················· |
| R | 5462 hCoV-19/England/QEUH-B133AC/2020\|EPI_ISL_641813\|20 | ···················· | ···················· | ····················· | ···················· | ···················· | ···················· |
| R | 5463 hCoV-19/England/QEUH-B12D45/2020\|EPI_ISL_641814\|20 | ···················· | ···················· | ····················· | ···················· | ···················· | ···················· |
| R | 5464 hCoV-19/England/QEUH-B1429F/2020\|EPI_ISL_641815\|20 | ···················· | ···················· | ····················· | ···················· | ···················· | ···················· |
| R | 5465 hCoV-19/England/QEUH-B139D7/2020\|EPI_ISL_641816\|20 | ···················· | ···················· | ····················· | ···················· | ···················· | ···················· |
| R | 5466 hCoV-19/England/QEUH-B1389E/2020\|EPI_ISL_641817\|20 | ···················· | ···················· | ····················· | ···················· | ···················· | ···················· |
| R | 5467 hCoV-19/England/QEUH-B139AA/2020\|EPI_ISL_641818\|20 | ···················· | ···················· | ····················· | ···················· | ···················· | ···················· |
| R | 5468 hCoV-19/England/QEUH-B0F7C5/2020\|EPI_ISL_641819\|20 | ···················· | ···················· | ····················· | ···················· | ···················· | ···················· |
| R | 5469 hCoV-19/England/QEUH-B0F701/2020\|EPI_ISL_641820\|20 | ···················· | ···················· | ····················· | ···················· | ···················· | ···················· |
| R | 5470 hCoV-19/England/QEUH-B0F103/2020\|EPI_ISL_641821\|20 | ···················· | ···················· | ····················· | ···················· | ···················· | ···················· |
| R | 5471 hCoV-19/England/QEUH-B0F84A/2020\|EPI_ISL_641823\|20 | ····G··············· | ···················· | ····················· | ···················· | ···················· | ···················· |
| R | 5472 hCoV-19/England/QEUH-B0F77A/2020\|EPI_ISL_641824\|20 | ···················· | ···················· | ····················· | ···················· | ···················· | ···················· |
| R | 5473 hCoV-19/England/QEUH-B145E1/2020\|EPI_ISL_641825\|20 | ···················· | ···················· | ····················· | ···················· | ···················· | ···················· |
| R | 5474 hCoV-19/England/QEUH-B139F5/2020\|EPI_ISL_641826\|20 | ···················· | ···················· | ····················· | ···················· | ···················· | ···················· |
| R | 5475 hCoV-19/England/QEUH-B14059/2020\|EPI_ISL_641828\|20 | ···················· | ···················· | ····················· | ···················· | ···················· | ···················· |
| R | 5476 hCoV-19/England/QEUH-B10776/2020\|EPI_ISL_641829\|20 | ···················· | ···················· | ····················· | ···················· | ···················· | ···················· |
| R | 5477 hCoV-19/England/QEUH-B13A6B/2020\|EPI_ISL_641830\|20 | ···················· | ···················· | ····················· | ···················· | ···················· | ···················· |
| R | 5478 hCoV-19/England/QEUH-B13B3B/2020\|EPI_ISL_641831\|20 | ···················· | ···················· | ····················· | ···················· | ···················· | ···················· |
| R | 5479 hCoV-19/England/QEUH-B13A89/2020\|EPI_ISL_641832\|20 | ···················· | ···················· | ····················· | ···················· | ···················· | ···················· |
| R | 5480 hCoV-19/England/QEUH-B13D80/2020\|EPI_ISL_641833\|20 | ···················· | ···················· | ····················· | ···················· | ···················· | ···················· |
| R | 5481 hCoV-19/England/QEUH-B13DAE/2020\|EPI_ISL_641834\|20 | ···················· | ···················· | ····················· | ···················· | ···················· | ···················· |
| R | 5482 hCoV-19/England/QEUH-B14086/2020\|EPI_ISL_641835\|20 | ···················· | ···················· | ····················· | ···················· | ···················· | ···················· |
| R | 5483 hCoV-19/England/QEUH-B13D62/2020\|EPI_ISL_641836\|20 | ···················· | ···················· | ····················· | ···················· | ···················· | ···················· |
| R | 5484 hCoV-19/England/QEUH-B107A3/2020\|EPI_ISL_641837\|20 | ···················· | ···················· | ····················· | ···················· | ···················· | ···················· |
| R | 5485 hCoV-19/England/QEUH-B142DB/2020\|EPI_ISL_641838\|20 | ···················· | ···················· | ····················· | ···················· | ···················· | ···················· |
| R | 5486 hCoV-19/England/QEUH-B13DBD/2020\|EPI_ISL_641839\|20 | ···················· | ···················· | ····················· | ···················· | ···················· | ···················· |
| R | 5487 hCoV-19/England/QEUH-B13A2F/2020\|EPI_ISL_641841\|20 | ···················· | ···················· | ····················· | ···················· | ···················· | ···················· |
| R | 5488 hCoV-19/England/QEUH-B13BFF/2020\|EPI_ISL_641842\|20 | ···················· | ···················· | ····················· | ···················· | ···················· | ···················· |
| R | 5489 hCoV-19/England/QEUH-B13E05/2020\|EPI_ISL_641843\|20 | ···················· | ···················· | ····················· | ···················· | ···················· | ···················· |
| R | 5490 hCoV-19/England/QEUH-B105E5/2020\|EPI_ISL_641844\|20 | ···················· | ···················· | ····················· | ···················· | ···················· | ···················· |
| R | 5491 hCoV-19/England/QEUH-B13F4E/2020\|EPI_ISL_641845\|20 | ···················· | ···················· | ····················· | ···················· | ···················· | ···················· |
| R | 5492 hCoV-19/England/QEUH-B141CF/2020\|EPI_ISL_641846\|20 | ···················· | ···················· | ····················· | ···················· | ···················· | ···················· |
| R | 5493 hCoV-19/England/QEUH-B107EF/2020\|EPI_ISL_641848\|20 | ···················· | ·A·················· | ····················· | ···················· | ···················· | ···················· |
| R | 5494 hCoV-19/England/QEUH-B14271/2020\|EPI_ISL_641849\|20 | ···················· | ···················· | ····················· | ···················· | ···················· | ···················· |
| R | 5495 hCoV-19/England/QEUH-B12C93/2020\|EPI_ISL_641851\|20 | ···················· | ···················· | ····················· | ···················· | ···················· | ···················· |
| R | 5496 hCoV-19/England/QEUH-B14208/2020\|EPI_ISL_641852\|20 | ···················· | ···················· | ····················· | ···················· | ···················· | ···················· |
| R | 5497 hCoV-19/England/QEUH-B12CEE/2020\|EPI_ISL_641853\|20 | ···················· | ···················· | ····················· | ···················· | ···················· | ···················· |
| R | 5498 hCoV-19/England/QEUH-B10758/2020\|EPI_ISL_641854\|20 | ···················· | ·A·················· | ····················· | ···················· | ···················· | ···················· |
| R | 5499 hCoV-19/England/QEUH-B1347C/2020\|EPI_ISL_641855\|20 | ···················· | ···················· | ····················· | ···················· | ···················· | ···················· |
| R | 5500 hCoV-19/England/QEUH-B0F859/2020\|EPI_ISL_641856\|20 | ···················· | ···················· | ····················· | ···················· | ···················· | ···················· |
| R | 5501 hCoV-19/England/QEUH-B13B77/2020\|EPI_ISL_641857\|20 | ···················· | ···················· | ····················· | ···················· | ···················· | ···················· |
| R | 5502 hCoV-19/England/QEUH-B106C4/2020\|EPI_ISL_641858\|20 | ···················· | ···················· | ····················· | ···················· | ···················· | ···················· |
| R | 5503 hCoV-19/England/QEUH-B0F710/2020\|EPI_ISL_641859\|20 | ···················· | ···················· | ····················· | ···················· | ···················· | ···················· |
| R | 5504 hCoV-19/England/QEUH-B106B5/2020\|EPI_ISL_641860\|20 | ···················· | ···················· | ····················· | ···················· | ···················· | ···················· |

| | | | | | | | | |
|---|---|---|---|---|---|---|---|---|
| R | 5505 | hCoV-19/England/QEUH-B13719/2020\|EPI_ISL_641861\|20 | .................... | .................... | .................... | .................... | .................... | .................... |
| R | 5506 | hCoV-19/England/QEUH-B14323/2020\|EPI_ISL_641863\|20 | .................... | .................... | .................... | .................... | .................... | .................... |
| R | 5507 | hCoV-19/England/QEUH-B13A4D/2020\|EPI_ISL_641864\|20 | .................... | .................... | .................... | .................... | .................... | .................... |
| R | 5508 | hCoV-19/England/QEUH-B13C92/2020\|EPI_ISL_641865\|20 | .................... | .................... | .................... | .................... | .................... | .................... |
| R | 5509 | hCoV-19/England/QEUH-B10819/2020\|EPI_ISL_641866\|20 | .................... | .................... | .................... | .................... | .................... | .................... |
| R | 5510 | hCoV-19/England/QEUH-B13A5C/2020\|EPI_ISL_641867\|20 | .................... | .................... | .................... | .................... | .................... | .................... |
| R | 5511 | hCoV-19/England/QEUH-B141B0/2020\|EPI_ISL_641868\|20 | .................... | .................... | .................... | .................... | .................... | .................... |
| R | 5512 | hCoV-19/England/QEUH-B0F112/2020\|EPI_ISL_641870\|20 | .................... | .................... | .................... | .................... | .................... | .................... |
| R | 5513 | hCoV-19/England/QEUH-B13C47/2020\|EPI_ISL_641871\|20 | .................... | .................... | .................... | .................... | .................... | .................... |
| R | 5514 | hCoV-19/England/QEUH-B0F81D/2020\|EPI_ISL_641872\|20 | .................... | .................... | .................... | .................... | .................... | .................... |
| R | 5515 | hCoV-19/England/QEUH-B1344F/2020\|EPI_ISL_641873\|20 | .................... | .................... | .................... | .................... | .................... | .................... |
| R | 5516 | hCoV-19/England/QEUH-B0F0D9/2020\|EPI_ISL_641874\|20 | .................... | .................... | .................... | .................... | .................... | .................... |
| R | 5517 | hCoV-19/England/QEUH-B1072B/2020\|EPI_ISL_641875\|20 | .................... | .................... | .................... | .................... | .................... | .................... |
| R | 5518 | hCoV-19/England/QEUH-B0F0F7/2020\|EPI_ISL_641876\|20 | .................... | .................... | .................... | .................... | .................... | .................... |
| R | 5519 | hCoV-19/England/QEUH-B13764/2020\|EPI_ISL_641877\|20 | .................... | .................... | .................... | .................... | .................... | .................... |
| R | 5520 | hCoV-19/England/QEUH-B12F4F/2020\|EPI_ISL_641878\|20 | .................... | .................... | .................... | .................... | ......T............. | .................... |
| R | 5521 | hCoV-19/England/QEUH-B13870/2020\|EPI_ISL_641879\|20 | .................... | .................... | .................... | .................... | .................... | .................... |
| R | 5522 | hCoV-19/England/QEUH-B1411A/2020\|EPI_ISL_641880\|20 | .................... | .................... | .................... | .................... | .................... | .................... |
| R | 5523 | hCoV-19/England/QEUH-B134E5/2020\|EPI_ISL_641882\|20 | .................... | .................... | .................... | .................... | .................... | .................... |
| R | 5524 | hCoV-19/England/QEUH-B13C1A/2020\|EPI_ISL_641883\|20 | .................... | .................... | .................... | .................... | .................... | .................... |
| R | 5525 | hCoV-19/England/QEUH-B12D63/2020\|EPI_ISL_641884\|20 | .................... | .................... | .................... | .................... | .................... | .................... |
| R | 5526 | hCoV-19/England/QEUH-B13816/2020\|EPI_ISL_641885\|20 | .................... | .................... | .................... | .................... | .................... | .................... |
| R | 5527 | hCoV-19/England/QEUH-B0F7E3/2020\|EPI_ISL_641886\|20 | .................... | .................... | .................... | .................... | .................... | .................... |
| R | 5528 | hCoV-19/England/QEUH-B13DF9/2020\|EPI_ISL_641887\|20 | .................... | .................... | .................... | .................... | .................... | .................... |
| R | 5529 | hCoV-19/England/QEUH-B13DDB/2020\|EPI_ISL_641888\|20 | .................... | .................... | .................... | .................... | .................... | .................... |
| R | 5530 | hCoV-19/England/QEUH-B144B7/2020\|EPI_ISL_641889\|20 | .................... | .................... | .................... | .................... | .................... | .................... |
| R | 5531 | hCoV-19/England/QEUH-B0F789/2020\|EPI_ISL_641890\|20 | .................... | .................... | .................... | .................... | .................... | .................... |
| R | 5532 | hCoV-19/England/QEUH-B10749/2020\|EPI_ISL_641891\|20 | .................... | .................... | .................... | .................... | .................... | .................... |
| R | 5533 | hCoV-19/England/QEUH-B1453C/2020\|EPI_ISL_641892\|20 | .................... | .................... | .................... | .................... | .................... | .................... |
| R | 5534 | hCoV-19/England/QEUH-B139E6/2020\|EPI_ISL_641893\|20 | .................... | .................... | .................... | .................... | .................... | .................... |
| R | 5535 | hCoV-19/England/QEUH-B136D0/2020\|EPI_ISL_641894\|20 | .................... | .................... | .................... | .................... | .................... | .................... |
| R | 5536 | hCoV-19/England/QEUH-B10837/2020\|EPI_ISL_641895\|20 | .................... | .................... | .................... | .................... | .................... | .................... |
| R | 5537 | hCoV-19/England/QEUH-B13C56/2020\|EPI_ISL_641896\|20 | .................... | .................... | .................... | .................... | .................... | .................... |
| R | 5538 | hCoV-19/England/QEUH-B13C65/2020\|EPI_ISL_641897\|20 | .................... | .................... | .................... | .................... | .................... | .................... |
| R | 5539 | hCoV-19/England/QEUH-B13C0B/2020\|EPI_ISL_641898\|20 | .................... | .................... | .................... | .................... | .................... | .................... |
| R | 5540 | hCoV-19/England/QEUH-B13940/2020\|EPI_ISL_641899\|20 | .................... | .................... | .................... | .................... | .................... | .................... |
| R | 5541 | hCoV-19/Scotland/QEUH-AD0306/2020\|EPI_ISL_644040\|2 | .................... | .................... | .................... | .................... | .................... | .................... |
| R | 5542 | hCoV-19/Scotland/QEUH-AD0421/2020\|EPI_ISL_644041\|2 | .................... | .................... | .................... | .................... | .................... | .................... |
| R | 5543 | hCoV-19/Scotland/QEUH-AD02CD/2020\|EPI_ISL_644044\|2 | .................... | .................... | .................... | .................... | .................... | .................... |
| R | 5544 | hCoV-19/Scotland/QEUH-AD2A5A/2020\|EPI_ISL_644045\|2 | .................... | .................... | .................... | .................... | .................... | .................... |
| R | 5545 | hCoV-19/Scotland/QEUH-AD070A/2020\|EPI_ISL_644046\|2 | .................... | .................... | .................... | .................... | .................... | .................... |
| R | 5546 | hCoV-19/Scotland/QEUH-AD26FC/2020\|EPI_ISL_644047\|2 | .................... | .................... | .................... | .................... | .................... | .................... |
| R | 5547 | hCoV-19/Scotland/QEUH-AD4AB2/2020\|EPI_ISL_644048\|2 | .................... | .................... | .................... | .................... | .................... | .................... |
| R | 5548 | hCoV-19/Scotland/QEUH-AD4A67/2020\|EPI_ISL_644049\|2 | .................... | .................... | .................... | .................... | .................... | .................... |
| R | 5549 | hCoV-19/Scotland/QEUH-AD62C7/2020\|EPI_ISL_644050\|2 | .................... | .................... | .................... | .................... | .................... | .................... |
| R | 5550 | hCoV-19/Scotland/QEUH-AD02DC/2020\|EPI_ISL_644051\|2 | .................... | .................... | .................... | .................... | .................... | .................... |
| R | 5551 | hCoV-19/England/MILK-AC940D/2020\|EPI_ISL_644052\|20 | .................... | .................... | .................... | .................... | ......T............. | .................... |
| R | 5552 | hCoV-19/England/MILK-AC77E8/2020\|EPI_ISL_644053\|20 | .................... | .................... | .................... | .................... | .................... | .................... |
| R | 5553 | hCoV-19/England/MILK-AC7557/2020\|EPI_ISL_644054\|20 | .................... | .................... | .................... | .................... | ......T............. | .................... |
| R | 5554 | hCoV-19/England/MILK-AC7566/2020\|EPI_ISL_644055\|20 | .................... | .................... | .................... | .................... | .................... | .................... |
| R | 5555 | hCoV-19/England/MILK-AC9388/2020\|EPI_ISL_644056\|20 | .................... | .................... | .................... | .................... | .................... | .................... |
| R | 5556 | hCoV-19/England/MILK-AC7575/2020\|EPI_ISL_644057\|20 | .................... | .................... | .................... | .................... | .................... | .................... |
| R | 5557 | hCoV-19/England/MILK-AC7548/2020\|EPI_ISL_644058\|20 | .................... | .................... | .................... | .................... | .................... | .................... |
| R | 5558 | hCoV-19/England/MILK-AC7A85/2020\|EPI_ISL_644059\|20 | .................... | .................... | .................... | .................... | .................... | .................... |
| R | 5559 | hCoV-19/England/MILK-AC7821/2020\|EPI_ISL_644061\|20 | .................... | .................... | .................... | .................... | .................... | .................... |
| R | 5560 | hCoV-19/England/MILK-AC7812/2020\|EPI_ISL_644063\|20 | .................... | .................... | .................... | .................... | .................... | .................... |
| R | 5561 | hCoV-19/England/MILK-ACD7D3/2020\|EPI_ISL_644064\|20 | .................... | .................... | .................... | .................... | ......T............. | .................... |
| R | 5562 | hCoV-19/England/MILK-ACD7B5/2020\|EPI_ISL_644066\|20 | .................... | .................... | .................... | .................... | .................... | .................... |
| R | 5563 | hCoV-19/England/MILK-ACDB7D/2020\|EPI_ISL_644067\|20 | .................... | .................... | .................... | .................... | .................... | .................... |
| R | 5564 | hCoV-19/England/MILK-AC77CA/2020\|EPI_ISL_644068\|20 | .................... | .................... | .................... | .................... | .................... | .................... |
| R | 5565 | hCoV-19/England/MILK-ACEC97/2020\|EPI_ISL_644069\|20 | .................... | .................... | .................... | .................... | ......T............. | .................... |
| R | 5566 | hCoV-19/England/MILK-ACE24A/2020\|EPI_ISL_644071\|20 | .................... | .................... | .................... | .................... | .................... | .................... |
| R | 5567 | hCoV-19/England/MILK-ACCA44/2020\|EPI_ISL_644072\|20 | .................... | .................... | .................... | .................... | ......T............. | .................... |
| R | 5568 | hCoV-19/England/MILK-ACC2E2/2020\|EPI_ISL_644073\|20 | .................... | .................... | .................... | .................... | .................... | .................... |
| R | 5569 | hCoV-19/England/MILK-ACED58/2020\|EPI_ISL_644074\|20 | .................... | .................... | .................... | .................... | .................... | .................... |
| R | 5570 | hCoV-19/England/MILK-ACE927/2020\|EPI_ISL_644075\|20 | .................... | .................... | .................... | .................... | .................... | .................... |
| R | 5571 | hCoV-19/England/MILK-ACC9CF/2020\|EPI_ISL_644076\|20 | .................... | .................... | .................... | .................... | .................... | .................... |
| R | 5572 | hCoV-19/England/MILK-ACC31C/2020\|EPI_ISL_644077\|20 | .................... | .................... | .................... | .................... | .................... | .................... |
| R | 5573 | hCoV-19/England/MILK-ACC2A6/2020\|EPI_ISL_644078\|20 | .................... | .................... | .................... | .................... | .................... | .................... |
| R | 5574 | hCoV-19/England/MILK-ACE550/2020\|EPI_ISL_644079\|20 | .................... | .................... | .................... | .................... | .................... | .................... |
| R | 5575 | hCoV-19/England/MILK-ACE277/2020\|EPI_ISL_644081\|20 | .................... | .................... | .................... | .................... | ......T............. | .................... |
| R | 5576 | hCoV-19/England/MILK-ACE541/2020\|EPI_ISL_644082\|20 | .................... | .................... | .................... | .................... | .................... | .................... |
| R | 5577 | hCoV-19/England/MILK-ACE75A/2020\|EPI_ISL_644083\|20 | .................... | .................... | .................... | .................... | .................... | .................... |
| R | 5578 | hCoV-19/England/MILK-ACEE37/2020\|EPI_ISL_644084\|20 | .................... | .................... | .................... | .................... | .................... | .................... |
| R | 5579 | hCoV-19/England/MILK-ACE7F0/2020\|EPI_ISL_644085\|20 | .................... | .................... | .................... | .................... | .................... | .................... |
| R | 5580 | hCoV-19/England/MILK-AB7754/2020\|EPI_ISL_644087\|20 | .................... | .................... | .................... | .................... | .................... | .................... |
| R | 5581 | hCoV-19/England/MILK-AB829E/2020\|EPI_ISL_644088\|20 | .................... | .................... | .................... | .................... | ......T............. | .................... |
| R | 5582 | hCoV-19/England/MILK-AB8489/2020\|EPI_ISL_644089\|20 | .................... | .................... | .................... | .................... | .................... | .................... |
| R | 5583 | hCoV-19/England/MILK-AB8647/2020\|EPI_ISL_644090\|20 | .................... | .................... | .................... | .................... | .................... | .................... |
| R | 5584 | hCoV-19/England/MILK-AB789D/2020\|EPI_ISL_644091\|20 | .................... | .................... | .................... | .................... | .................... | .................... |

| | | | | | | | |
|---|---|---|---|---|---|---|---|
| R | 5585 hCoV-19/Scotland/QEUH-AAE802/2020\|EPI_ISL_644094\|2 | ···················· | ···················· | ····················· | ···················· | ···················· | ···················· |
| R | 5586 hCoV-19/England/ALDP-AA0958/2020\|EPI_ISL_644095\|20 | ···················· | ···················· | ····················· | ···················· | ···················· | ···················· |
| R | 5587 hCoV-19/England/ALDP-AA2965/2020\|EPI_ISL_644096\|20 | ···················· | ···················· | ····················· | ···················· | ···················· | ···················· |
| R | 5588 hCoV-19/England/ALDP-AA2956/2020\|EPI_ISL_644097\|20 | ···················· | ···················· | ····················· | ···················· | ···················· | ···················· |
| R | 5589 hCoV-19/England/CAMC-B3607F/2020\|EPI_ISL_645370\|20 | ···················· | ···················· | ····················· | ···················· | ···················· | ···················· |
| R | 5590 hCoV-19/England/CAMC-B363B2/2020\|EPI_ISL_645371\|20 | ···················· | ···················· | ····················· | ···················· | ···················· | ···················· |
| R | 5591 hCoV-19/England/CAMC-B372B4/2020\|EPI_ISL_645372\|20 | ···················· | ···················· | ····················· | ···················· | ······T············· | ···················· |
| R | 5592 hCoV-19/England/CAMC-B3717B/2020\|EPI_ISL_645373\|20 | ···················· | ···················· | ····················· | ···················· | ···················· | ···················· |
| R | 5593 hCoV-19/England/MILK-B3D2BE/2020\|EPI_ISL_645374\|20 | ···················· | ···················· | ····················· | ···················· | ···················· | ···················· |
| R | 5594 hCoV-19/England/CAMC-B371B7/2020\|EPI_ISL_645376\|20 | ···················· | ···················· | ····················· | ···················· | ···················· | ···················· |
| R | 5595 hCoV-19/England/CAMC-B36358/2020\|EPI_ISL_645379\|20 | ···················· | ···················· | ····················· | ···················· | ···················· | ···················· |
| R | 5596 hCoV-19/England/CAMC-B372E1/2020\|EPI_ISL_645381\|20 | ···················· | ···················· | ····················· | ···················· | ···················· | ···················· |
| R | 5597 hCoV-19/England/MILK-B3D1B1/2020\|EPI_ISL_645383\|20 | ···················· | ···················· | ····················· | ···················· | ···················· | ···················· |
| R | 5598 hCoV-19/England/CAMC-B36A53/2020\|EPI_ISL_645386\|20 | ···················· | ···················· | ····················· | ···················· | ···················· | ···················· |
| R | 5599 hCoV-19/England/CAMC-B3730C/2020\|EPI_ISL_645387\|20 | ···················· | ···················· | ····················· | ···················· | ···················· | ···················· |
| R | 5600 hCoV-19/England/CAMC-B35ED0/2020\|EPI_ISL_645389\|20 | ···················· | ···················· | ····················· | ···················· | ···················· | ···················· |
| R | 5601 hCoV-19/England/MILK-B3D4F4/2020\|EPI_ISL_645391\|20 | ···················· | ···················· | ····················· | ···················· | ···················· | ···················· |
| R | 5602 hCoV-19/England/CAMC-B37296/2020\|EPI_ISL_645392\|20 | ···················· | ···················· | ····················· | ···················· | ······T············· | ···················· |
| R | 5603 hCoV-19/England/MILK-B3D290/2020\|EPI_ISL_645393\|20 | ···················· | ··········Y········· | ····················· | ···················· | ···················· | ···················· |
| R | 5604 hCoV-19/England/MILK-B3D4A9/2020\|EPI_ISL_645395\|20 | ···················· | ···················· | ····················· | ···················· | ···················· | ···················· |
| R | 5605 hCoV-19/England/CAMC-B35E3A/2020\|EPI_ISL_645397\|20 | ···················· | ···················· | ····················· | ···················· | ···················· | ···················· |
| R | 5606 hCoV-19/England/CAMC-B375F7/2020\|EPI_ISL_645398\|20 | ···················· | ···················· | ····················· | ···················· | ···················· | ···················· |
| R | 5607 hCoV-19/England/MILK-B3D579/2020\|EPI_ISL_645399\|20 | ···················· | ···················· | ····················· | ···················· | ···················· | ···················· |
| R | 5608 hCoV-19/England/CAMC-B36974/2020\|EPI_ISL_645410\|20 | ···················· | ···················· | ····················· | ···················· | ···················· | ···················· |
| R | 5609 hCoV-19/England/CAMC-B3681D/2020\|EPI_ISL_645411\|20 | ···················· | ···················· | ····················· | ···················· | ···················· | ···················· |
| R | 5610 hCoV-19/England/CAMC-B370F6/2020\|EPI_ISL_645416\|20 | ···················· | ···················· | ····················· | ···················· | ···················· | ···················· |
| R | 5611 hCoV-19/England/CAMC-B36929/2020\|EPI_ISL_645417\|20 | ···················· | ···················· | ····················· | ···················· | ···················· | ···················· |
| R | 5612 hCoV-19/England/ALDP-B3EC91/2020\|EPI_ISL_645418\|20 | ···················· | ···················· | ····················· | ···················· | ···················· | ···················· |
| R | 5613 hCoV-19/England/CAMC-B37454/2020\|EPI_ISL_645419\|20 | ···················· | ···················· | ····················· | ···················· | ···················· | ···················· |
| R | 5614 hCoV-19/England/CAMC-B374FA/2020\|EPI_ISL_645420\|20 | ···················· | ···················· | ····················· | ···················· | ···················· | ···················· |
| R | 5615 hCoV-19/England/MILK-B3D6C1/2020\|EPI_ISL_645421\|20 | ···················· | ···················· | ····················· | ···················· | ······T············· | ···················· |
| R | 5616 hCoV-19/England/CAMC-B365F8/2020\|EPI_ISL_645422\|20 | ···················· | ·A·················· | ····················· | ···················· | ···················· | ···A················ |
| R | 5617 hCoV-19/England/MILK-B3D272/2020\|EPI_ISL_645424\|20 | ···················· | ···················· | ····················· | ···················· | ···················· | ···················· |
| R | 5618 hCoV-19/England/CAMC-B364A0/2020\|EPI_ISL_645425\|20 | ···················· | ···················· | ····················· | ···················· | ···················· | ···················· |
| R | 5619 hCoV-19/England/CAMC-B371E4/2020\|EPI_ISL_645426\|20 | ···················· | ···················· | ····················· | ···················· | ···················· | ···················· |
| R | 5620 hCoV-19/England/ALDP-B24DD7/2020\|EPI_ISL_646740\|20 | ···················· | ···················· | ····················· | ···················· | ···················· | ···················· |
| R | 5621 hCoV-19/England/ALDP-B250BF/2020\|EPI_ISL_646741\|20 | ···················· | ···················· | ····················· | ···················· | ···················· | ···················· |
| R | 5622 hCoV-19/England/QEUH-B2B9EB/2020\|EPI_ISL_646742\|20 | ···················· | ·A·················· | ····················· | ···················· | ···················· | ···················· |
| R | 5623 hCoV-19/England/ALDP-B24DB9/2020\|EPI_ISL_646744\|20 | ···················· | ···················· | ····················· | ···················· | ···················· | ···················· |
| R | 5624 hCoV-19/England/ALDP-B24E98/2020\|EPI_ISL_646747\|20 | ···················· | ···················· | ····················· | ···················· | ···················· | ···················· |
| R | 5625 hCoV-19/England/QEUH-B2B4BD/2020\|EPI_ISL_646748\|20 | ···················· | ···················· | ····················· | ···················· | ···················· | ···················· |
| R | 5626 hCoV-19/England/ALDP-B24E01/2020\|EPI_ISL_646749\|20 | ···················· | ···················· | ····················· | ···················· | ···················· | ···················· |
| R | 5627 hCoV-19/England/ALDP-B25F85/2020\|EPI_ISL_646751\|20 | ···················· | ···················· | ····················· | ···················· | ···················· | ···················· |
| R | 5628 hCoV-19/England/ALDP-B263D3/2020\|EPI_ISL_646752\|20 | ···················· | ···················· | ····················· | ···················· | ···················· | ···················· |
| R | 5629 hCoV-19/England/ALDP-B24E2F/2020\|EPI_ISL_646753\|20 | ···················· | ···················· | ····················· | ···················· | ···················· | ···················· |
| R | 5630 hCoV-19/England/QEUH-B2C8DE/2020\|EPI_ISL_646754\|20 | ···················· | ···················· | ····················· | ···················· | ···················· | ···················· |
| R | 5631 hCoV-19/England/ALDP-B24D31/2020\|EPI_ISL_646755\|20 | ···················· | ·A·················· | ····················· | ···················· | ···················· | ···················· |
| R | 5632 hCoV-19/England/ALDP-B26151/2020\|EPI_ISL_646757\|20 | ···················· | ···················· | ····················· | ···················· | ···················· | ···················· |
| R | 5634 hCoV-19/Australia/VIC16249/2020\|EPI_ISL_640480\|202 | ···················· | ···················· | ····················· | ···················· | ···················· | ···················· |
| R | 5636 hCoV-19/Australia/VIC15982/2020\|EPI_ISL_640485\|202 | ···················· | ···················· | ····················· | ···················· | ···················· | ···················· |
| R | 5637 hCoV-19/Australia/VIC12176/2020\|EPI_ISL_640486\|202 | ···················· | ···················· | ····················· | ···················· | ···················· | ···················· |
| R | 5638 hCoV-19/Australia/VIC16265/2020\|EPI_ISL_640487\|202 | ···················· | ···················· | ····················· | ···················· | ···················· | ···················· |
| R | 5639 hCoV-19/Australia/VIC12766/2020\|EPI_ISL_640490\|202 | ···················· | ···················· | ····················· | ···················· | ···················· | ···················· |
| R | 5641 hCoV-19/Australia/VIC12331/2020\|EPI_ISL_640494\|202 | ···················· | ···················· | ····················· | ···················· | ···················· | ···················· |
| R | 5642 hCoV-19/Australia/VIC12390/2020\|EPI_ISL_640495\|202 | ···················· | ···················· | ····················· | ···················· | ···················· | ···················· |
| R | 5643 hCoV-19/Australia/VIC16510/2020\|EPI_ISL_640496\|202 | ···················· | ···················· | ····················· | ···················· | ···················· | ···················· |
| R | 5644 hCoV-19/Australia/VIC16113/2020\|EPI_ISL_640497\|202 | ···················· | ···················· | ····················· | ···················· | ···················· | ···················· |
| R | 5645 hCoV-19/Australia/VIC16127/2020\|EPI_ISL_640498\|202 | ···················· | ···················· | ····················· | ···················· | ···················· | ···················· |
| R | 5646 hCoV-19/Australia/VIC15927/2020\|EPI_ISL_640499\|202 | ···················· | ···················· | ····················· | ···················· | ···················· | ···················· |
| R | 5647 hCoV-19/Australia/VIC12748/2020\|EPI_ISL_640530\|202 | ···················· | ···················· | ····················· | ···················· | ···················· | ···················· |
| R | 5648 hCoV-19/Australia/VIC16108/2020\|EPI_ISL_640531\|202 | ···················· | ···················· | ····················· | ···················· | ···················· | ···················· |
| R | 5649 hCoV-19/England/QEUH-B137BF/2020\|EPI_ISL_641862\|20 | ···················· | ···················· | ····················· | ···················· | ···················· | ···················· |
| R | 5650 hCoV-19/Scotland/QEUH-AD0315/2020\|EPI_ISL_644000\|2 | ···················· | ···················· | ····················· | ···················· | ···················· | ···················· |
| R | 5651 hCoV-19/Scotland/QEUH-AD6388/2020\|EPI_ISL_644001\|2 | ···················· | ···················· | ····················· | ···················· | ···················· | ···················· |
| R | 5652 hCoV-19/Scotland/QEUH-AD2AB4/2020\|EPI_ISL_644002\|2 | ···················· | ···················· | ····················· | ···················· | ···················· | ···················· |
| R | 5653 hCoV-19/Scotland/QEUH-AD4AEF/2020\|EPI_ISL_644004\|2 | ···················· | ···················· | ····················· | ···················· | ···················· | ···················· |
| R | 5654 hCoV-19/Scotland/QEUH-AD2744/2020\|EPI_ISL_644005\|2 | ···················· | ···················· | ····················· | ···················· | ···················· | ···················· |
| R | 5655 hCoV-19/Scotland/QEUH-AD1E31/2020\|EPI_ISL_644009\|2 | ···················· | ···················· | ····················· | ···················· | ···················· | ···················· |
| R | 5656 hCoV-19/Scotland/QEUH-AD2AD2/2020\|EPI_ISL_644010\|2 | ···················· | ···················· | ····················· | ···················· | ···················· | ···················· |
| R | 5657 hCoV-19/Scotland/QEUH-AD4AD0/2020\|EPI_ISL_644011\|2 | ···················· | ···················· | ····················· | ···················· | ···················· | ···················· |
| R | 5658 hCoV-19/Scotland/QEUH-AD02AF/2020\|EPI_ISL_644012\|2 | ···················· | ···················· | ····················· | ···················· | ···················· | ···················· |
| R | 5659 hCoV-19/Scotland/QEUH-AD1D07/2020\|EPI_ISL_644013\|2 | ···················· | ···················· | ····················· | ···················· | ···················· | ···················· |
| R | 5660 hCoV-19/Scotland/QEUH-AD4AC1/2020\|EPI_ISL_644014\|2 | ···················· | ···················· | ····················· | ···················· | ···················· | ···················· |
| R | 5661 hCoV-19/Scotland/QEUH-AD2665/2020\|EPI_ISL_644016\|2 | ···················· | ···················· | ····················· | ···················· | ···················· | ···················· |
| R | 5662 hCoV-19/Scotland/QEUH-AD4B0A/2020\|EPI_ISL_644017\|2 | ···················· | ···················· | ····················· | ···················· | ···················· | ···················· |
| R | 5663 hCoV-19/Scotland/QEUH-AD2B1B/2020\|EPI_ISL_644018\|2 | ···················· | ···················· | ····················· | ···················· | ···················· | ···················· |
| R | 5664 hCoV-19/Scotland/QEUH-AD2AF0/2020\|EPI_ISL_644019\|2 | ·······T············ | ···················· | ····················· | ···················· | ···················· | ···················· |
| R | 5665 hCoV-19/Scotland/QEUH-AD1DDA/2020\|EPI_ISL_644020\|2 | ···················· | ···················· | ····················· | ···················· | ···················· | ···················· |
| R | 5666 hCoV-19/Scotland/QEUH-AD4A85/2020\|EPI_ISL_644021\|2 | ···················· | ···················· | ····················· | ···················· | ···················· | ···················· |
| R | 5667 hCoV-19/Scotland/QEUH-AD1D16/2020\|EPI_ISL_644023\|2 | ···················· | ···················· | ····················· | ···················· | ···················· | ···················· |

| | | | | | | | |
|---|---|---|---|---|---|---|---|
| R | 5668 hCoV-19/Scotland/QEUH-AD4B64/2020\|EPI_ISL_644024\|2 | ···················· | ···················· | ····················· | ···················· | ···················· | ···················· |
| R | 5669 hCoV-19/Scotland/QEUH-AD2A96/2020\|EPI_ISL_644025\|2 | ···················· | ···················· | ····················· | ···················· | ···················· | ···················· |
| R | 5670 hCoV-19/Scotland/QEUH-AD4B37/2020\|EPI_ISL_644026\|2 | ···················· | ···················· | ····················· | ···················· | ···················· | ···················· |
| R | 5671 hCoV-19/Scotland/QEUH-AD1E40/2020\|EPI_ISL_644027\|2 | ···················· | ···················· | ····················· | ···················· | ···················· | ···················· |
| R | 5672 hCoV-19/Scotland/QEUH-AD4B28/2020\|EPI_ISL_644028\|2 | ···················· | ···················· | ····················· | ···················· | ···················· | ···················· |
| R | 5673 hCoV-19/Scotland/QEUH-AD2708/2020\|EPI_ISL_644030\|2 | ···················· | ···················· | ····················· | ···················· | ···················· | ···················· |
| R | 5674 hCoV-19/Scotland/QEUH-AD06D0/2020\|EPI_ISL_644031\|2 | ···················· | ···················· | ····················· | ···················· | ···················· | ···················· |
| R | 5675 hCoV-19/Scotland/QEUH-AD1E9B/2020\|EPI_ISL_644032\|2 | ···················· | ···················· | ····················· | ···················· | ···················· | ···················· |
| R | 5676 hCoV-19/Scotland/QEUH-AD4B82/2020\|EPI_ISL_644034\|2 | ···················· | ···················· | ····················· | ···················· | ···················· | ···················· |
| R | 5677 hCoV-19/Scotland/QEUH-AD4B91/2020\|EPI_ISL_644035\|2 | ···················· | ···················· | ····················· | ···················· | ···················· | ···················· |
| R | 5678 hCoV-19/Scotland/QEUH-AD4AFE/2020\|EPI_ISL_644036\|2 | ···················· | ···················· | ····················· | ···················· | ···················· | ···················· |
| R | 5679 hCoV-19/Scotland/QEUH-AD4B73/2020\|EPI_ISL_644037\|2 | ···················· | ···················· | ····················· | ···················· | ···················· | ···················· |
| R | 5680 hCoV-19/Scotland/QEUH-AD1D52/2020\|EPI_ISL_644038\|2 | ···················· | ···················· | ····················· | ···················· | ···················· | ···················· |
| R | 5681 hCoV-19/Scotland/QEUH-AD1E04/2020\|EPI_ISL_644039\|2 | ···················· | ···················· | ····················· | ···················· | ···················· | ···················· |
| R | 5682 hCoV-19/England/CAMC-B369FC/2020\|EPI_ISL_645307\|20 | ···················· | ···················· | ····················· | ···················· | ···················· | ···················· |
| R | 5683 hCoV-19/England/MILK-B3D4C7/2020\|EPI_ISL_645308\|20 | ···················· | ···················· | ····················· | ···················· | ···················· | ···················· |
| R | 5684 hCoV-19/England/CAMC-B36938/2020\|EPI_ISL_645309\|20 | ···················· | ···················· | ····················· | ···················· | ···················· | ···················· |
| R | 5685 hCoV-19/England/CAMC-B35E2B/2020\|EPI_ISL_645310\|20 | ···················· | ···················· | ····················· | ···················· | ···················· | ···················· |
| R | 5686 hCoV-19/England/CAMC-B366D7/2020\|EPI_ISL_645311\|20 | ···················· | ···················· | ····················· | ···················· | ···················· | ···················· |
| R | 5687 hCoV-19/England/CAMC-B375D9/2020\|EPI_ISL_645312\|20 | ···················· | ···················· | ····················· | ···················· | ···················· | ···················· |
| R | 5688 hCoV-19/England/CAMC-B37366/2020\|EPI_ISL_645313\|20 | ···················· | ···················· | ····················· | ···················· | ······T············· | ···················· |
| R | 5689 hCoV-19/England/CAMC-B37490/2020\|EPI_ISL_645314\|20 | ···················· | ···················· | ····················· | ···················· | ···················· | ···················· |
| R | 5690 hCoV-19/England/CAMC-B375AC/2020\|EPI_ISL_645315\|20 | ···················· | ···················· | ····················· | ···················· | ···················· | ···················· |
| R | 5691 hCoV-19/England/CAMC-B3672F/2020\|EPI_ISL_645316\|20 | ···················· | ···················· | ····················· | ···················· | ···················· | ···················· |
| R | 5692 hCoV-19/England/CAMC-B360AC/2020\|EPI_ISL_645317\|20 | ···················· | ·A·················· | ····················· | ···················· | ···················· | ···················· |
| R | 5693 hCoV-19/England/CAMC-B367B6/2020\|EPI_ISL_645318\|20 | ···················· | ···················· | ····················· | ···················· | ···················· | ···················· |
| R | 5694 hCoV-19/England/MILK-B3D55B/2020\|EPI_ISL_645319\|20 | ···················· | ···················· | ····················· | ···················· | ···················· | ···················· |
| R | 5695 hCoV-19/England/CAMC-B3640A/2020\|EPI_ISL_645320\|20 | ···················· | ···················· | ····················· | ···················· | ······T············· | ···················· |
| R | 5696 hCoV-19/England/CAMC-B3721E/2020\|EPI_ISL_645321\|20 | ···················· | ···················· | ····················· | ···················· | ···················· | ···················· |
| R | 5697 hCoV-19/England/CAMC-B36570/2020\|EPI_ISL_645322\|20 | ···················· | ···················· | ····················· | ···················· | ···················· | ···················· |
| R | 5698 hCoV-19/England/CAMC-B35FFB/2020\|EPI_ISL_645323\|20 | ···················· | ···················· | ····················· | ···················· | ······T············· | ···················· |
| R | 5699 hCoV-19/England/MILK-B3D236/2020\|EPI_ISL_645324\|20 | ···················· | ···················· | ····················· | ···················· | ···················· | ···················· |
| R | 5700 hCoV-19/England/CAMC-B362B5/2020\|EPI_ISL_645325\|20 | ···················· | ···················· | ····················· | ···················· | ···················· | ···················· |
| R | 5701 hCoV-19/England/CAMC-B3622E/2020\|EPI_ISL_645326\|20 | ···················· | ···················· | ····················· | ···················· | ···················· | ···················· |
| R | 5702 hCoV-19/England/MILK-B3D4E5/2020\|EPI_ISL_645327\|20 | ···················· | ···················· | ····················· | ···················· | ···················· | ···················· |
| R | 5703 hCoV-19/England/CAMC-B361A9/2020\|EPI_ISL_645328\|20 | ···················· | ···················· | ····················· | ···················· | ···················· | ···················· |
| R | 5704 hCoV-19/England/MILK-B3D48B/2020\|EPI_ISL_645329\|20 | ···················· | ···················· | ····················· | ···················· | ···················· | ···················· |
| R | 5705 hCoV-19/England/CAMC-B35EEF/2020\|EPI_ISL_645331\|20 | ···················· | ···················· | ····················· | ···················· | ···················· | ···················· |
| R | 5706 hCoV-19/England/CAMC-B366C8/2020\|EPI_ISL_645332\|20 | ···················· | ···················· | ····················· | ···················· | ···················· | ···················· |
| R | 5707 hCoV-19/England/CAMC-B372C3/2020\|EPI_ISL_645333\|20 | ···················· | ···················· | ····················· | ···················· | ······T············· | ···················· |
| R | 5708 hCoV-19/England/CAMC-B37463/2020\|EPI_ISL_645334\|20 | ···················· | ···················· | ····················· | ···················· | ···················· | ···················· |
| R | 5709 hCoV-19/England/CAMC-B375E8/2020\|EPI_ISL_645335\|20 | ···················· | ···················· | ····················· | ···················· | ···················· | ···················· |
| R | 5710 hCoV-19/England/CAMC-B368D1/2020\|EPI_ISL_645337\|20 | ···················· | ···················· | ····················· | ···················· | ······T············· | ···················· |
| R | 5711 hCoV-19/England/CAMC-B36376/2020\|EPI_ISL_645338\|20 | ···················· | ···················· | ····················· | ···················· | ···················· | ···················· |
| R | 5712 hCoV-19/England/CAMC-B366B9/2020\|EPI_ISL_645339\|20 | ···················· | ···················· | ····················· | ···················· | ······T············· | ···················· |
| R | 5713 hCoV-19/England/CAMC-B3669B/2020\|EPI_ISL_645342\|20 | ···················· | ···················· | ····················· | ···················· | ···················· | ···················· |
| R | 5714 hCoV-19/England/CAMC-B37418/2020\|EPI_ISL_645343\|20 | ···················· | ···················· | ····················· | ···················· | ···················· | ···················· |
| R | 5715 hCoV-19/England/CAMC-B3616D/2020\|EPI_ISL_645344\|20 | ···················· | ···················· | ····················· | ···················· | ···················· | ···················· |
| R | 5716 hCoV-19/England/CAMC-B364BF/2020\|EPI_ISL_645346\|20 | ···················· | ···················· | ····················· | ···················· | ···················· | ···················· |
| R | 5717 hCoV-19/England/CAMC-B36491/2020\|EPI_ISL_645347\|20 | ···················· | ·A········A········· | ····················· | ···················· | ···················· | ···················· |
| R | 5718 hCoV-19/England/CAMC-B360F7/2020\|EPI_ISL_645349\|20 | ···················· | ···················· | ····················· | ···················· | ······T············· | ···················· |
| R | 5719 hCoV-19/England/CAMC-B3720F/2020\|EPI_ISL_645350\|20 | ···················· | ···················· | ····················· | ···················· | ···················· | ···················· |
| R | 5720 hCoV-19/England/CAMC-B36868/2020\|EPI_ISL_645351\|20 | ···················· | ···················· | ····················· | ···················· | ···················· | ···················· |
| R | 5721 hCoV-19/England/CAMC-B37348/2020\|EPI_ISL_645352\|20 | ···················· | ···················· | ····················· | ···················· | ···················· | ···················· |
| R | 5722 hCoV-19/England/CAMC-B36701/2020\|EPI_ISL_645353\|20 | ···················· | ···················· | ····················· | ···················· | ···················· | ···················· |
| R | 5723 hCoV-19/England/MILK-B3D6D0/2020\|EPI_ISL_645354\|20 | ···················· | ···················· | ····················· | ···················· | ···················· | ···················· |
| R | 5724 hCoV-19/England/CAMC-B37393/2020\|EPI_ISL_645355\|20 | ···················· | ···················· | ····················· | ···················· | ···················· | ···················· |
| R | 5725 hCoV-19/England/CAMC-B3718A/2020\|EPI_ISL_645356\|20 | ···················· | ···················· | ····················· | ···················· | ···················· | ···················· |
| R | 5726 hCoV-19/England/CAMC-B374AF/2020\|EPI_ISL_645357\|20 | ···················· | ···················· | ····················· | ···················· | ···················· | ···················· |
| R | 5727 hCoV-19/England/CAMC-B3673E/2020\|EPI_ISL_645358\|20 | ···················· | ·A·················· | ····················· | ···················· | ···················· | ···A················ |
| R | 5728 hCoV-19/England/CAMC-B36561/2020\|EPI_ISL_645359\|20 | ···················· | ···················· | ····················· | ···················· | ······T············· | ···················· |
| R | 5729 hCoV-19/England/MILK-B3D6A3/2020\|EPI_ISL_645360\|20 | ···················· | ···················· | ····················· | ···················· | ···················· | ···················· |
| R | 5730 hCoV-19/England/CAMC-B37384/2020\|EPI_ISL_645361\|20 | ···················· | ···················· | ····················· | ···················· | ···················· | ···················· |
| R | 5731 hCoV-19/England/ALDP-B3EF3E/2020\|EPI_ISL_645362\|20 | ···················· | ···················· | ····················· | ···················· | ···················· | ···················· |
| R | 5732 hCoV-19/England/CAMC-B37357/2020\|EPI_ISL_645363\|20 | ···················· | ···················· | ····················· | ···················· | ···················· | ···················· |
| R | 5733 hCoV-19/England/CAMC-B3713F/2020\|EPI_ISL_645364\|20 | ···················· | ···················· | ····················· | ···················· | ···················· | ···················· |
| R | 5734 hCoV-19/England/CAMC-B365BC/2020\|EPI_ISL_645366\|20 | ···················· | ···················· | ····················· | ···················· | ···················· | ···················· |
| R | 5735 hCoV-19/England/MILK-B3D4B8/2020\|EPI_ISL_645367\|20 | ···················· | ···················· | ····················· | ···················· | ···················· | ···················· |
| R | 5736 hCoV-19/England/ALDP-B3F128/2020\|EPI_ISL_645368\|20 | ···················· | ···················· | ····················· | ···················· | ···················· | ···················· |
| R | 5737 hCoV-19/England/MILK-B3D184/2020\|EPI_ISL_645369\|20 | ···················· | ·A·················· | ····················· | ···················· | ···················· | ···A················ |
| R | 5738 hCoV-19/England/ALDP-B2632E/2020\|EPI_ISL_646600\|20 | ···················· | ···················· | ····················· | ···················· | ···················· | ···················· |
| R | 5739 hCoV-19/England/ALDP-B26054/2020\|EPI_ISL_646601\|20 | ···················· | ···················· | ····················· | ···················· | ···················· | ···················· |
| R | 5740 hCoV-19/England/QEUH-B2B7A5/2020\|EPI_ISL_646602\|20 | ···················· | ···················· | ····················· | ···················· | ···················· | ···················· |
| R | 5741 hCoV-19/England/ALDP-B24DAA/2020\|EPI_ISL_646603\|20 | ···················· | ···················· | ····················· | ···················· | ···················· | ···················· |
| R | 5742 hCoV-19/England/QEUH-B2B453/2020\|EPI_ISL_646604\|20 | ···················· | ···················· | ····················· | ···················· | ···················· | ···················· |
| R | 5743 hCoV-19/England/ALDP-B26106/2020\|EPI_ISL_646605\|20 | ···················· | ···················· | ····················· | ···················· | ···················· | ···················· |
| R | 5744 hCoV-19/England/ALDP-B252AA/2020\|EPI_ISL_646606\|20 | ···················· | ·A·················· | ····················· | ···················· | ···················· | ···················· |
| R | 5745 hCoV-19/England/ALDP-B25EF1/2020\|EPI_ISL_646607\|20 | ···················· | ···················· | ····················· | ···················· | ···················· | ···················· |
| R | 5746 hCoV-19/England/QEUH-B2C917/2020\|EPI_ISL_646608\|20 | ···················· | ···················· | ····················· | ···················· | ···················· | ···················· |
| R | 5747 hCoV-19/England/QEUH-B2B58D/2020\|EPI_ISL_646609\|20 | ···················· | ···················· | ····················· | ···················· | ···················· | ···················· |

| | | | | | | | |
|---|---|---|---|---|---|---|---|
| R | 5748 hCoV-19/England/ALDP-B26063/2020\|EPI_ISL_646610\|20 | ···················· | ···················· | ···················· | ···················· | ···················· | ···················· |
| R | 5749 hCoV-19/England/ALDP-B2629A/2020\|EPI_ISL_646612\|20 | ···················· | ···················· | ···················· | ···················· | ···················· | ···················· |
| R | 5750 hCoV-19/England/ALDP-B260BE/2020\|EPI_ISL_646613\|20 | ···················· | ···················· | ···················· | ···················· | ···················· | ···················· |
| R | 5751 hCoV-19/England/ALDP-B260CD/2020\|EPI_ISL_646614\|20 | ···················· | ···················· | ···················· | ···················· | ···················· | ···················· |
| R | 5752 hCoV-19/England/QEUH-B2B3FC/2020\|EPI_ISL_646615\|20 | ···················· | ···················· | ···················· | ···················· | ···················· | ···················· |
| R | 5753 hCoV-19/England/ALDP-B2619D/2020\|EPI_ISL_646616\|20 | ···················· | ···················· | ···················· | ···················· | ···················· | ···················· |
| R | 5754 hCoV-19/England/QEUH-B2B532/2020\|EPI_ISL_646617\|20 | ···················· | ···················· | ···················· | ···················· | ······T············· | ···················· |
| R | 5755 hCoV-19/England/ALDP-B24F4A/2020\|EPI_ISL_646618\|20 | ···················· | ···················· | ···················· | ···················· | ···················· | ···················· |
| R | 5756 hCoV-19/England/QEUH-B2B981/2020\|EPI_ISL_646619\|20 | ···················· | ···················· | ···················· | ···················· | ···················· | ···················· |
| R | 5757 hCoV-19/England/QEUH-B2B417/2020\|EPI_ISL_646620\|20 | ···················· | ···················· | ···················· | ···················· | ···················· | ···················· |
| R | 5758 hCoV-19/England/QEUH-B2B945/2020\|EPI_ISL_646622\|20 | ···················· | ···················· | ···················· | ···················· | ···················· | ···················· |
| R | 5759 hCoV-19/England/ALDP-B250EC/2020\|EPI_ISL_646623\|20 | ···················· | ···················· | ···················· | ···················· | ···················· | ···················· |
| R | 5760 hCoV-19/England/QEUH-B2CAF6/2020\|EPI_ISL_646624\|20 | ···················· | ···················· | ···················· | ···················· | ···················· | ···················· |
| R | 5761 hCoV-19/England/ALDP-B24ED4/2020\|EPI_ISL_646625\|20 | ···················· | ···················· | ···················· | ···················· | ···················· | ···················· |
| R | 5762 hCoV-19/England/QEUH-B2C980/2020\|EPI_ISL_646637\|20 | ···················· | ···················· | ···················· | ···················· | ···················· | ···················· |
| R | 5763 hCoV-19/England/ALDP-B24F95/2020\|EPI_ISL_646638\|20 | ···················· | ·A·················· | ···················· | ···················· | ···················· | ···················· |
| R | 5764 hCoV-19/England/ALDP-B24D7D/2020\|EPI_ISL_646639\|20 | ···················· | ···················· | ···················· | ···················· | ···················· | ···················· |
| R | 5765 hCoV-19/England/ALDP-B261F7/2020\|EPI_ISL_646640\|20 | ···················· | ···················· | ···················· | ···················· | ···················· | ···················· |
| R | 5766 hCoV-19/England/QEUH-B2B57E/2020\|EPI_ISL_646641\|20 | ···················· | ···················· | ···················· | ···················· | ···················· | ···················· |
| R | 5767 hCoV-19/England/ALDP-B262B8/2020\|EPI_ISL_646642\|20 | ···················· | ···················· | ···················· | ···················· | ···················· | ···················· |
| R | 5768 hCoV-19/England/QEUH-B2B426/2020\|EPI_ISL_646643\|20 | ···················· | ···················· | ···················· | ···················· | ···················· | ···A················ |
| R | 5769 hCoV-19/England/QEUH-B2C9AE/2020\|EPI_ISL_646645\|20 | ···················· | ·A·················· | ···················· | ···················· | ···················· | ···················· |
| R | 5770 hCoV-19/England/QEUH-B2B4DB/2020\|EPI_ISL_646646\|20 | ···················· | ···················· | ···················· | ···················· | ···················· | ···················· |
| R | 5771 hCoV-19/England/QEUH-B2B6A8/2020\|EPI_ISL_646647\|20 | ···················· | ···················· | ···················· | ···················· | ···················· | ···················· |
| R | 5772 hCoV-19/England/ALDP-B2519E/2020\|EPI_ISL_646648\|20 | ···················· | ···················· | ···················· | ···················· | ······T············· | ···················· |
| R | 5773 hCoV-19/England/QEUH-B2B884/2020\|EPI_ISL_646649\|20 | ···················· | ···················· | ···················· | ···················· | ···················· | ···················· |
| R | 5774 hCoV-19/England/QEUH-B2B347/2020\|EPI_ISL_646650\|20 | ···················· | ···················· | ···················· | ···················· | ····T··············· | ···················· |
| R | 5775 hCoV-19/England/ALDP-B26115/2020\|EPI_ISL_646651\|20 | ···················· | ···················· | ···················· | ···················· | ···················· | ···················· |
| R | 5776 hCoV-19/England/ALDP-B25EB5/2020\|EPI_ISL_646652\|20 | ···················· | ···················· | ···················· | ···················· | ···················· | ···················· |
| R | 5777 hCoV-19/England/ALDP-B24F2C/2020\|EPI_ISL_646653\|20 | ···················· | ···················· | ···················· | ···················· | ···················· | ···················· |
| R | 5778 hCoV-19/England/ALDP-B262C7/2020\|EPI_ISL_646655\|20 | ···················· | ···················· | ···················· | ···················· | ···················· | ···················· |
| R | 5779 hCoV-19/England/QEUH-B2BC97/2020\|EPI_ISL_646656\|20 | ···················· | ···················· | ···················· | ···················· | ···················· | ···A················ |
| R | 5780 hCoV-19/England/QEUH-B2C908/2020\|EPI_ISL_646657\|20 | ···················· | ···················· | ···················· | ···················· | ···················· | ···················· |
| R | 5781 hCoV-19/England/ALDP-B24F3B/2020\|EPI_ISL_646658\|20 | ···················· | ·A·················· | ···················· | ···················· | ···················· | ···A················ |
| R | 5782 hCoV-19/England/QEUH-B2BABB/2020\|EPI_ISL_646659\|20 | ···················· | ···················· | ···················· | ···················· | ···················· | ···················· |
| R | 5783 hCoV-19/England/ALDP-B250DD/2020\|EPI_ISL_646660\|20 | ···················· | ···················· | ···················· | ···················· | ···················· | ···················· |
| R | 5784 hCoV-19/England/QEUH-B2B408/2020\|EPI_ISL_646661\|20 | ···················· | ···················· | ···················· | ···················· | ···················· | ···················· |
| R | 5785 hCoV-19/England/ALDP-B250CE/2020\|EPI_ISL_646662\|20 | ···················· | ···················· | ···················· | ···················· | ···················· | ···················· |
| R | 5786 hCoV-19/England/QEUH-B2B839/2020\|EPI_ISL_646663\|20 | ···················· | ···················· | ···················· | ···················· | ···················· | ···················· |
| R | 5787 hCoV-19/England/ALDP-B263A6/2020\|EPI_ISL_646664\|20 | ···················· | ···················· | ···················· | ···················· | ···················· | ···················· |
| R | 5788 hCoV-19/England/ALDP-B24D22/2020\|EPI_ISL_646665\|20 | ···················· | ···················· | ···················· | ···················· | ···················· | ···················· |
| R | 5789 hCoV-19/England/ALDP-B24FA4/2020\|EPI_ISL_646666\|20 | ···················· | ···················· | ···················· | ···················· | ···················· | ···················· |
| R | 5790 hCoV-19/England/QEUH-B2B7C3/2020\|EPI_ISL_646667\|20 | ···················· | ···················· | ···················· | ···················· | ···················· | ···················· |
| R | 5791 hCoV-19/England/ALDP-B26081/2020\|EPI_ISL_646668\|20 | ···················· | ·A·················· | ···················· | ···················· | ···················· | ···A················ |
| R | 5792 hCoV-19/England/QEUH-B2BB03/2020\|EPI_ISL_646669\|20 | ···················· | ···················· | ···················· | ···················· | ···················· | ···················· |
| R | 5793 hCoV-19/England/QEUH-B2B769/2020\|EPI_ISL_646670\|20 | ···················· | ···················· | ···················· | ···················· | ···················· | ···················· |
| R | 5794 hCoV-19/England/ALDP-B262F4/2020\|EPI_ISL_646671\|20 | ···················· | ···················· | ···················· | ···················· | ···················· | ···················· |
| R | 5795 hCoV-19/England/QEUH-B2B611/2020\|EPI_ISL_646672\|20 | ···················· | ···················· | ···················· | ···················· | ···················· | ···················· |
| R | 5796 hCoV-19/England/ALDP-B260FA/2020\|EPI_ISL_646673\|20 | ···················· | ···················· | ···················· | ···················· | ···················· | ···················· |
| R | 5797 hCoV-19/England/ALDP-B26124/2020\|EPI_ISL_646675\|20 | ···················· | ···················· | ···················· | ···················· | ···················· | ···················· |
| R | 5798 hCoV-19/England/QEUH-B2B796/2020\|EPI_ISL_646677\|20 | ···················· | ···················· | ···················· | ···················· | ···················· | ···················· |
| R | 5799 hCoV-19/England/QEUH-B2B383/2020\|EPI_ISL_646680\|20 | ···················· | ···················· | ···················· | ···················· | ···················· | ···················· |
| R | 5800 hCoV-19/England/QEUH-B2B63F/2020\|EPI_ISL_646681\|20 | ···················· | ···················· | ···················· | ···················· | ···················· | ···················· |
| R | 5801 hCoV-19/England/QEUH-B2BAE8/2020\|EPI_ISL_646682\|20 | ···················· | ···················· | ···················· | ···················· | ······T············· | ···················· |
| R | 5802 hCoV-19/England/ALDP-B2617F/2020\|EPI_ISL_646683\|20 | ···················· | ···················· | ···················· | ···················· | ···················· | ···················· |
| R | 5803 hCoV-19/England/ALDP-B26300/2020\|EPI_ISL_646684\|20 | ···················· | ···················· | ···················· | ···················· | ···················· | ···················· |
| R | 5804 hCoV-19/England/QEUH-B2C8CF/2020\|EPI_ISL_646685\|20 | ···················· | ···················· | ···················· | ···················· | ···················· | ···················· |
| R | 5805 hCoV-19/England/ALDP-B261CA/2020\|EPI_ISL_646686\|20 | ···················· | ···················· | ···················· | ···················· | ···················· | ···················· |
| R | 5806 hCoV-19/England/ALDP-B2526E/2020\|EPI_ISL_646687\|20 | ···················· | ···················· | ···················· | ···················· | ···················· | ···················· |
| R | 5807 hCoV-19/England/QEUH-B2B392/2020\|EPI_ISL_646688\|20 | ···················· | ···················· | ···················· | ···················· | ···················· | ···················· |
| R | 5808 hCoV-19/England/ALDP-B26009/2020\|EPI_ISL_646689\|20 | ···················· | ···················· | ···················· | ···················· | ···················· | ···················· |
| R | 5809 hCoV-19/England/QEUH-B2B5C9/2020\|EPI_ISL_646690\|20 | ···················· | ···················· | ···················· | ···················· | ···················· | ···················· |
| R | 5810 hCoV-19/England/ALDP-B26160/2020\|EPI_ISL_646691\|20 | ···················· | ···················· | ···················· | ···················· | ···················· | ···················· |
| R | 5811 hCoV-19/England/ALDP-B252F5/2020\|EPI_ISL_646692\|20 | ···················· | ···················· | ···················· | ···················· | ···················· | ···················· |
| R | 5812 hCoV-19/England/ALDP-B24D9B/2020\|EPI_ISL_646693\|20 | ···················· | ···················· | ···················· | ···················· | ···················· | ···················· |
| R | 5813 hCoV-19/England/QEUH-B2BB21/2020\|EPI_ISL_646694\|20 | ···················· | ···················· | ···················· | ···················· | ···················· | ···················· |
| R | 5814 hCoV-19/England/ALDP-B25125/2020\|EPI_ISL_646695\|20 | ···················· | ·A·················· | ···················· | ···················· | ···················· | ···················· |
| R | 5815 hCoV-19/England/ALDP-B2633D/2020\|EPI_ISL_646696\|20 | ···················· | ·A·················· | ···················· | ···················· | ···················· | ···A················ |
| R | 5816 hCoV-19/England/ALDP-B25E97/2020\|EPI_ISL_646697\|20 | ···················· | ···················· | ···················· | ···················· | ···················· | ···················· |
| R | 5817 hCoV-19/England/ALDP-B24FC2/2020\|EPI_ISL_646698\|20 | ···················· | ···················· | ···················· | ···················· | ···················· | ···················· |
| R | 5818 hCoV-19/Wales/CAMC-AEAEEA/2020\|EPI_ISL_647900\|2020 | ···················· | ···················· | ···················· | ···················· | ···················· | ···················· |
| R | 5819 hCoV-19/Wales/CAMC-AEAF05/2020\|EPI_ISL_647901\|2020 | ···················· | ···················· | ···················· | ···················· | ···················· | ···················· |
| R | 5820 hCoV-19/Wales/QEUH-AE4A73/2020\|EPI_ISL_647902\|2020 | ···················· | ···················· | ···················· | ···················· | ···················· | ···················· |
| R | 5821 hCoV-19/Wales/QEUH-AE4AFB/2020\|EPI_ISL_647904\|2020 | ···················· | ···················· | ···················· | ···················· | ···················· | ···················· |
| R | 5822 hCoV-19/Wales/QEUH-AE519B/2020\|EPI_ISL_647905\|2020 | ···················· | ···················· | ···················· | ···················· | ···················· | ···················· |
| R | 5823 hCoV-19/Wales/CAMC-AEAF14/2020\|EPI_ISL_647906\|2020 | ···················· | ···················· | ···················· | ···················· | ···················· | ···················· |
| R | 5824 hCoV-19/Wales/QEUH-AE4AEC/2020\|EPI_ISL_647907\|2020 | ···················· | ···················· | ···················· | ···················· | ···················· | ···················· |
| R | 5825 hCoV-19/Wales/CAMC-AEAF41/2020\|EPI_ISL_647908\|2020 | ···················· | ···················· | ···················· | ···················· | ···················· | ···················· |
| R | 5826 hCoV-19/Wales/CAMC-AEAF23/2020\|EPI_ISL_647909\|2020 | ···················· | ···················· | ···················· | ···················· | ···················· | ···················· |
| R | 5827 hCoV-19/Wales/QEUH-AE4BE9/2020\|EPI_ISL_647910\|2020 | ···················· | ···················· | ···················· | ···················· | ···················· | ···················· |

| | | | | | | | |
|---|---|---|---|---|---|---|---|
| R | 5828 hCoV-19/Wales/QEUH-AE4BDA/2020\|EPI_ISL_647911\|2020 | ···················· | ···················· | ····················· | ···················· | ···················· | ···················· |
| R | 5829 hCoV-19/Wales/QEUH-AE4B25/2020\|EPI_ISL_647912\|2020 | ···················· | ···················· | ····················· | ···················· | ···················· | ···················· |
| R | 5830 hCoV-19/Wales/CAMC-AEAEF9/2020\|EPI_ISL_647913\|2020 | ···················· | ···················· | ····················· | ···················· | ···················· | ···················· |
| R | 5831 hCoV-19/Wales/QEUH-AE4B8F/2020\|EPI_ISL_647914\|2020 | ···················· | ···················· | ····················· | ···················· | ···················· | ···················· |
| R | 5832 hCoV-19/Wales/QEUH-AE4ABF/2020\|EPI_ISL_647915\|2020 | ···················· | ···················· | ····················· | ···················· | ···················· | ···················· |
| R | 5833 hCoV-19/Wales/CAMC-AEAF32/2020\|EPI_ISL_647916\|2020 | ···················· | ···················· | ····················· | ···················· | ···················· | ···················· |
| R | 5834 hCoV-19/Wales/QEUH-AE525C/2020\|EPI_ISL_647917\|2020 | ···················· | ···················· | ····················· | ···················· | ···················· | ···················· |
| R | 5835 hCoV-19/Wales/QEUH-AE517D/2020\|EPI_ISL_647918\|2020 | ···················· | ···················· | ····················· | ···················· | ···················· | ···················· |
| R | 5836 hCoV-19/Wales/QEUH-AE4AA0/2020\|EPI_ISL_647919\|2020 | ···················· | ···················· | ····················· | ···················· | ···················· | ···················· |
| R | 5837 hCoV-19/Wales/QEUH-AE515F/2020\|EPI_ISL_647920\|2020 | ···················· | ···················· | ····················· | ···················· | ···················· | ···················· |
| R | 5838 hCoV-19/Wales/QEUH-AD58E4/2020\|EPI_ISL_647921\|2020 | ···················· | ···················· | ····················· | ···················· | ···················· | ···················· |
| R | 5839 hCoV-19/Wales/QEUH-AD4302/2020\|EPI_ISL_647922\|2020 | ···················· | ···················· | ····················· | ···················· | ···················· | ···················· |
| R | 5840 hCoV-19/Wales/QEUH-AD57BA/2020\|EPI_ISL_647923\|2020 | ···················· | ···················· | ····················· | ···················· | ···················· | ···················· |
| R | 5841 hCoV-19/Wales/QEUH-AD5BEB/2020\|EPI_ISL_647924\|2020 | ···················· | ···················· | ····················· | ···················· | ···················· | ···················· |
| R | 5842 hCoV-19/Wales/QEUH-AD4320/2020\|EPI_ISL_647925\|2020 | ···················· | ···················· | ····················· | ···················· | ···················· | ···················· |
| R | 5843 hCoV-19/Wales/QEUH-AD42AB/2020\|EPI_ISL_647926\|2020 | ···················· | ···················· | ····················· | ···················· | ···················· | ···················· |
| R | 5844 hCoV-19/Wales/QEUH-AD428D/2020\|EPI_ISL_647927\|2020 | ···················· | ···················· | ····················· | ···················· | ···················· | ···················· |
| R | 5845 hCoV-19/Wales/QEUH-AD4241/2020\|EPI_ISL_647928\|2020 | ···················· | ···················· | ····················· | ···················· | ···················· | ···················· |
| R | 5846 hCoV-19/Wales/QEUH-AD42C9/2020\|EPI_ISL_647929\|2020 | ···················· | ···················· | ····················· | ···················· | ···················· | ···················· |
| R | 5847 hCoV-19/Wales/QEUH-AD4205/2020\|EPI_ISL_647930\|2020 | ···················· | ···················· | ····················· | ···················· | ···················· | ···················· |
| R | 5848 hCoV-19/Wales/QEUH-AD5802/2020\|EPI_ISL_647931\|2020 | ···················· | ···················· | ····················· | ···················· | ···················· | ···················· |
| R | 5849 hCoV-19/Wales/QEUH-AD57C9/2020\|EPI_ISL_647932\|2020 | ···················· | ···················· | ····················· | ···················· | ···················· | ···················· |
| R | 5850 hCoV-19/Wales/QEUH-AD3F2D/2020\|EPI_ISL_647933\|2020 | ···················· | ···················· | ····················· | ···················· | ···················· | ···················· |
| R | 5851 hCoV-19/Wales/QEUH-AD427E/2020\|EPI_ISL_647934\|2020 | ···················· | ···················· | ····················· | ···················· | ···················· | ···················· |
| R | 5852 hCoV-19/Wales/QEUH-AD42F6/2020\|EPI_ISL_647935\|2020 | ···················· | ···················· | ····················· | ···················· | ···················· | ···················· |
| R | 5853 hCoV-19/Wales/QEUH-AD5811/2020\|EPI_ISL_647936\|2020 | ···················· | ···················· | ····················· | ···················· | ···················· | ···················· |
| R | 5854 hCoV-19/Wales/QEUH-AD5899/2020\|EPI_ISL_647937\|2020 | ···················· | ···················· | ····················· | ···················· | ···················· | ···················· |
| R | 5855 hCoV-19/Wales/QEUH-AD5723/2020\|EPI_ISL_647938\|2020 | ···················· | ···················· | ····················· | ···················· | ···················· | ···················· |
| R | 5856 hCoV-19/Wales/QEUH-AD426F/2020\|EPI_ISL_647939\|2020 | ···················· | ···················· | ····················· | ···················· | ···················· | ···················· |
| R | 5857 hCoV-19/Wales/QEUH-AD577E/2020\|EPI_ISL_647940\|2020 | ···················· | ···················· | ····················· | ···················· | ···················· | ···················· |
| R | 5858 hCoV-19/Wales/QEUH-AD3EF3/2020\|EPI_ISL_647941\|2020 | ···················· | ···················· | ····················· | ···················· | ···················· | ···················· |
| R | 5859 hCoV-19/Wales/QEUH-AD41F9/2020\|EPI_ISL_647942\|2020 | ···················· | ···················· | ····················· | ···················· | ···················· | ···················· |
| R | 5860 hCoV-19/Wales/QEUH-AD4153/2020\|EPI_ISL_647943\|2020 | ···················· | ···················· | ····················· | ···················· | ···················· | ···················· |
| R | 5861 hCoV-19/Wales/QEUH-AD32CA/2020\|EPI_ISL_647944\|2020 | ···················· | ···················· | ····················· | ···················· | ···················· | ···················· |
| R | 5862 hCoV-19/Wales/QEUH-AD1208/2020\|EPI_ISL_647945\|2020 | ···················· | ···················· | ····················· | ···················· | ···················· | ···················· |
| R | 5863 hCoV-19/Wales/QEUH-AD2823/2020\|EPI_ISL_647946\|2020 | ···················· | ···················· | ····················· | ···················· | ···················· | ···················· |
| R | 5864 hCoV-19/Wales/QEUH-AD3330/2020\|EPI_ISL_647947\|2020 | ···················· | ···················· | ····················· | ···················· | ···················· | ···················· |
| R | 5865 hCoV-19/Wales/QEUH-AD335E/2020\|EPI_ISL_647948\|2020 | ···················· | ···················· | ····················· | ···················· | ···················· | ···················· |
| R | 5866 hCoV-19/Wales/QEUH-AD338B/2020\|EPI_ISL_647949\|2020 | ···················· | ···················· | ····················· | ···················· | ···················· | ···················· |
| R | 5867 hCoV-19/Wales/QEUH-AD5D9A/2020\|EPI_ISL_647951\|2020 | ···················· | ···················· | ····················· | ···················· | ··A················· | ···················· |
| R | 5868 hCoV-19/Wales/QEUH-AD10E0/2020\|EPI_ISL_647953\|2020 | ···················· | ···················· | ····················· | ···················· | ···················· | ···················· |
| R | 5869 hCoV-19/Wales/QEUH-AD591E/2020\|EPI_ISL_647954\|2020 | ···················· | ···················· | ····················· | ···················· | ···················· | ···················· |
| R | 5870 hCoV-19/Wales/QEUH-AD0DEA/2020\|EPI_ISL_647955\|2020 | ···················· | ···················· | ····················· | ···················· | ···················· | ···················· |
| R | 5871 hCoV-19/Wales/QEUH-AD14C6/2020\|EPI_ISL_647956\|2020 | ···················· | ···················· | ····················· | ···················· | ···················· | ···················· |
| R | 5872 hCoV-19/Wales/QEUH-AD3479/2020\|EPI_ISL_647968\|2020 | ···················· | ···················· | ····················· | ···················· | ···················· | ···················· |
| R | 5873 hCoV-19/Zimbabwe/ZW-29/2020\|EPI_ISL_647978\|2020-03 | ···················· | ···················· | ····················· | ···················· | ···················· | ···················· |
| R | 5874 hCoV-19/USA/RI-CDC-8135/2020\|EPI_ISL_647985\|2020-0 | ···················· | ···················· | ····················· | ···················· | ···················· | ···················· |
| R | 5875 hCoV-19/USA/IN-CDC-4982/2020\|EPI_ISL_647986\|2020-0 | ···················· | ···················· | ····················· | ···················· | ···················· | ···················· |
| R | 5876 hCoV-19/USA/GA-CDC-5478/2020\|EPI_ISL_647987\|2020-0 | ···················· | ···················· | ····················· | ···················· | ···················· | ···················· |
| R | 5877 hCoV-19/USA/GA-CDC-5479/2020\|EPI_ISL_647988\|2020-0 | ···················· | ···················· | ····················· | ···················· | ···················· | ···················· |
| R | 5878 hCoV-19/USA/GA-CDC-5527/2020\|EPI_ISL_647989\|2020-0 | ···················· | ···················· | ····················· | ···················· | ···················· | ···················· |
| R | 5879 hCoV-19/USA/IA-CDC-4042/2020\|EPI_ISL_647990\|2020-0 | ···················· | ···················· | ····················· | ···················· | ···················· | ···················· |
| R | 5880 hCoV-19/USA/IA-CDC-4043/2020\|EPI_ISL_647991\|2020-0 | ···················· | ···················· | ····················· | ···················· | ···················· | ···················· |
| R | 5881 hCoV-19/USA/IA-CDC-4046/2020\|EPI_ISL_647992\|2020-0 | ···················· | ···················· | ····················· | ···················· | ···················· | ···················· |
| R | 5882 hCoV-19/USA/IA-CDC-4047/2020\|EPI_ISL_647993\|2020-0 | ···················· | ···················· | ····················· | ···················· | ···················· | ···················· |
| R | 5883 hCoV-19/USA/IA-CDC-4048/2020\|EPI_ISL_647994\|2020-0 | ···················· | ···················· | ····················· | ···················· | ···················· | ···················· |
| R | 5884 hCoV-19/USA/IA-CDC-4049/2020\|EPI_ISL_647995\|2020-0 | ···················· | ···················· | ····················· | ···················· | ···················· | ···················· |
| R | 5885 hCoV-19/USA/IA-CDC-4050/2020\|EPI_ISL_647996\|2020-0 | ···················· | ···················· | ····················· | ···················· | ···················· | ···················· |
| R | 5886 hCoV-19/USA/IA-CDC-4051/2020\|EPI_ISL_647997\|2020-0 | ···················· | ···················· | ····················· | ···················· | ···················· | ···················· |
| R | 5887 hCoV-19/USA/IA-CDC-4052/2020\|EPI_ISL_647998\|2020-0 | ···················· | ···················· | ····················· | ···················· | ···················· | ···················· |
| R | 5888 hCoV-19/USA/IA-CDC-4053/2020\|EPI_ISL_647999\|2020-0 | ···················· | ···················· | ····················· | ···················· | ···················· | ···················· |
| R | 5889 hCoV-19/England/QEUH-B0DB70/2020\|EPI_ISL_642116\|20 | ···················· | ···················· | ····················· | ···················· | ···················· | ···················· |
| R | 5890 hCoV-19/England/QEUH-B0EA27/2020\|EPI_ISL_642117\|20 | ···················· | ···················· | ····················· | ···················· | ···················· | ···················· |
| R | 5891 hCoV-19/England/QEUH-B0E043/2020\|EPI_ISL_642119\|20 | ···················· | ···················· | ····················· | ···················· | ···················· | ···················· |
| R | 5892 hCoV-19/England/QEUH-B0E1D7/2020\|EPI_ISL_642120\|20 | ···················· | ···················· | ····················· | ···················· | ···················· | ···················· |
| R | 5893 hCoV-19/England/QEUH-B0DC04/2020\|EPI_ISL_642122\|20 | ···················· | ·A·················· | ····················· | ···················· | ···················· | ···················· |
| R | 5894 hCoV-19/England/QEUH-B0DBCB/2020\|EPI_ISL_642123\|20 | ···················· | ···················· | ····················· | ···················· | ···················· | ···················· |
| R | 5895 hCoV-19/England/QEUH-B0E33B/2020\|EPI_ISL_642124\|20 | ···················· | ···················· | ····················· | ···················· | ···················· | ···················· |
| R | 5896 hCoV-19/England/QEUH-B0DDD4/2020\|EPI_ISL_642125\|20 | ···················· | ···················· | ····················· | ···················· | ···················· | ···················· |
| R | 5897 hCoV-19/England/QEUH-B0E2C5/2020\|EPI_ISL_642126\|20 | ···················· | ···················· | ····················· | ···················· | ···················· | ···················· |
| R | 5898 hCoV-19/England/QEUH-B0DB34/2020\|EPI_ISL_642127\|20 | ···················· | ···················· | ····················· | ···················· | ···················· | ···················· |
| R | 5899 hCoV-19/England/QEUH-B0E32C/2020\|EPI_ISL_642128\|20 | ···················· | ···················· | ····················· | ···················· | ···················· | ···················· |
| R | 5900 hCoV-19/England/QEUH-B0F868/2020\|EPI_ISL_642129\|20 | ···················· | ···················· | ····················· | ···················· | ···················· | ···················· |
| R | 5901 hCoV-19/England/QEUH-B0F394/2020\|EPI_ISL_642130\|20 | ···················· | ···················· | ····················· | ···················· | ···················· | ···················· |
| R | 5902 hCoV-19/England/QEUH-B0F30D/2020\|EPI_ISL_642131\|20 | ···················· | ···················· | ····················· | ···················· | ···················· | ···················· |
| R | 5903 hCoV-19/England/QEUH-B0F32B/2020\|EPI_ISL_642132\|20 | ···················· | ···················· | ····················· | ···················· | ···················· | ···················· |
| R | 5904 hCoV-19/England/QEUH-B0DBE9/2020\|EPI_ISL_642135\|20 | ···················· | ···················· | ····················· | ···················· | ···················· | ···················· |
| R | 5905 hCoV-19/England/QEUH-B0E0F8/2020\|EPI_ISL_642136\|20 | ···················· | ···················· | ····················· | ···················· | ···················· | ···················· |
| R | 5906 hCoV-19/England/QEUH-B0E993/2020\|EPI_ISL_642137\|20 | ···················· | ···················· | ····················· | ···················· | ···················· | ···················· |
| R | 5907 hCoV-19/England/CAMC-B0B5FC/2020\|EPI_ISL_643447\|20 | ···················· | ···················· | ····················· | ···················· | ···················· | ···················· |

| | | | | | | | | |
|---|---|---|---|---|---|---|---|---|
| R | 5908 hCoV-19/England/CAMC-B0B170/2020\|EPI_ISL_643449\|20 | ···················· | ···················· | ···················· | ···················· | ·····T·············· | ···················· | |
| R | 5909 hCoV-19/England/CAMC-B0BA75/2020\|EPI_ISL_643450\|20 | ···················· | ···················· | ···················· | ···················· | ···················· | ···················· | |
| R | 5910 hCoV-19/England/CAMC-B07B1C/2020\|EPI_ISL_643451\|20 | ···················· | ···················· | ···················· | ···················· | ···················· | ···················· | |
| R | 5911 hCoV-19/England/CAMC-B0C8F2/2020\|EPI_ISL_643452\|20 | ···················· | ···················· | ···················· | ···················· | ···················· | ···················· | |
| R | 5912 hCoV-19/England/CAMC-B0AEC5/2020\|EPI_ISL_643454\|20 | ···················· | ···················· | ···················· | ···················· | ···················· | ···················· | |
| R | 5913 hCoV-19/England/CAMC-B0CAB0/2020\|EPI_ISL_643455\|20 | ···················· | ···················· | ···················· | ···················· | ···················· | ···················· | |
| R | 5914 hCoV-19/England/CAMC-B0B055/2020\|EPI_ISL_643456\|20 | ···················· | ···················· | ···················· | ···················· | ···················· | ···················· | |
| R | 5915 hCoV-19/England/CAMC-B0C458/2020\|EPI_ISL_643457\|20 | ···················· | ···················· | ···················· | ···················· | ···················· | ···················· | |
| R | 5916 hCoV-19/England/CAMC-B0B44A/2020\|EPI_ISL_643459\|20 | ···················· | ···················· | ···················· | ···················· | ·····T·············· | ···················· | |
| R | 5917 hCoV-19/England/CAMC-B0CBEA/2020\|EPI_ISL_643460\|20 | ···················· | ···················· | ···················· | ···················· | ···················· | ···················· | |
| R | 5918 hCoV-19/England/CAMC-B0B26E/2020\|EPI_ISL_643461\|20 | ···················· | ···················· | ···················· | ···················· | ···················· | ···················· | |
| R | 5919 hCoV-19/England/CAMC-B080A3/2020\|EPI_ISL_643462\|20 | ···················· | ···················· | ···················· | ···················· | ···················· | ···················· | |
| R | 5920 hCoV-19/England/CAMC-B0B40E/2020\|EPI_ISL_643463\|20 | ···················· | ···················· | ···················· | ···················· | ···················· | ···················· | |
| R | 5921 hCoV-19/England/CAMC-B08234/2020\|EPI_ISL_643464\|20 | ···················· | ···················· | ···················· | ···················· | ···················· | ···················· | |
| R | 5922 hCoV-19/England/CAMC-B0854A/2020\|EPI_ISL_643465\|20 | ···················· | ···················· | ···················· | ···················· | ···················· | ···················· | |
| R | 5923 hCoV-19/England/CAMC-B0AF4A/2020\|EPI_ISL_643466\|20 | ···················· | ···················· | ···················· | ···················· | ···················· | ···················· | |
| R | 5924 hCoV-19/England/CAMC-B0B161/2020\|EPI_ISL_643467\|20 | ···················· | ···················· | ···················· | ···················· | ·····T·············· | ···················· | |
| R | 5925 hCoV-19/England/CAMC-B0B37A/2020\|EPI_ISL_643468\|20 | ···················· | ···················· | ···················· | ···················· | ···················· | ···················· | |
| R | 5926 hCoV-19/England/CAMC-B0C8B6/2020\|EPI_ISL_643500\|20 | ···················· | ···················· | ···················· | ···················· | ···················· | ···················· | |
| R | 5931 hCoV-19/Zimbabwe/ZW-6965/2020\|EPI_ISL_644790\|2020 | ···················· | ···················· | ···················· | ···················· | ···················· | ···················· | |
| R | 5936 hCoV-19/Zimbabwe/ZW-6978/2020\|EPI_ISL_644795\|2020- | ···················· | ···················· | ···················· | ···················· | ···················· | ···················· | |
| R | 5939 hCoV-19/Zimbabwe/ZW-6981/2020\|EPI_ISL_644798\|2020- | ···················· | ···················· | ···················· | ···················· | ···················· | ···················· | |
| R | 5941 hCoV-19/Zimbabwe/ZW-6989/2020\|EPI_ISL_644800\|2020- | ···················· | ···················· | ···················· | ···················· | ···················· | ···················· | |
| R | 5944 hCoV-19/Zimbabwe/ZW-6992/2020\|EPI_ISL_644803\|2020- | ···················· | ···················· | ···················· | ···················· | N·N················· | ···················· | |
| R | 5945 hCoV-19/Zimbabwe/ZW-6993/2020\|EPI_ISL_644804\|2020- | ···················· | ···················· | ···················· | ···················· | ···················· | ···················· | |
| R | 5946 hCoV-19/Zimbabwe/ZW-6996/2020\|EPI_ISL_644805\|2020- | ···················· | ···················· | ···················· | ···················· | ···················· | ···················· | |
| R | 5947 hCoV-19/Zimbabwe/ZW-6997/2020\|EPI_ISL_644806\|2020- | ···················· | ···················· | ···················· | ···················· | NNN················· | ···················· | 1 |
| R | 5948 hCoV-19/Zimbabwe/ZW-6998/2020\|EPI_ISL_644807\|2020- | ···················· | ···················· | ···················· | ···················· | NNN················· | ···················· | 1 |
| R | 5950 hCoV-19/Zimbabwe/ZW-7001/2020\|EPI_ISL_644809\|2020- | ···················· | ···················· | ···················· | ···················· | NNN················· | ···················· | 1 |
| R | 5953 hCoV-19/Zimbabwe/ZW-9494/2020\|EPI_ISL_644812\|2020- | ···················· | ·A·················· | ···················· | ···················· | ···················· | ···················· | |
| R | 5957 hCoV-19/Zimbabwe/ZW-9527/2020\|EPI_ISL_644816\|2020- | ···················· | ·A·················· | ···················· | ···················· | ···················· | ···················· | |
| R | 5959 hCoV-19/Zimbabwe/ZW-9947/2020\|EPI_ISL_644818\|2020- | ···················· | ···················· | ···················· | ···················· | ···················· | ···················· | |
| R | 5961 hCoV-19/Zimbabwe/ZW-9953/2020\|EPI_ISL_644820\|2020- | ···················· | ···················· | ···················· | ···················· | ···················· | ···················· | |
| R | 5963 hCoV-19/USA/VA-DCLS-1648/2020\|EPI_ISL_644825\|2020- | ···················· | ···················· | ···················· | ···················· | ···················· | ···················· | |
| R | 5964 hCoV-19/USA/VA-DCLS-1651/2020\|EPI_ISL_644826\|2020- | ···················· | ···················· | ···················· | ···················· | ···················· | ···················· | |
| R | 5965 hCoV-19/USA/VA-DCLS-1652/2020\|EPI_ISL_644827\|2020- | ···················· | ···················· | ···················· | ···················· | ···················· | ···················· | |
| R | 5966 hCoV-19/USA/VA-DCLS-1657/2020\|EPI_ISL_644829\|2020- | ···················· | ···················· | ···················· | ···················· | ···················· | ···················· | |
| R | 5967 hCoV-19/USA/VA-DCLS-1658/2020\|EPI_ISL_644830\|2020- | ···················· | ···················· | ···················· | ···················· | ···················· | ···················· | |
| R | 5968 hCoV-19/USA/VA-DCLS-1659/2020\|EPI_ISL_644831\|2020- | ···················· | ···················· | ···················· | ···················· | ···················· | ···················· | |
| R | 5969 hCoV-19/USA/VA-DCLS-1661/2020\|EPI_ISL_644832\|2020- | ···················· | ···················· | ···················· | ···················· | ···················· | ···················· | |
| R | 5970 hCoV-19/England/MILK-B3D500/2020\|EPI_ISL_645300\|20 | ···················· | ···················· | ···················· | ···················· | ···················· | ···················· | |
| R | 5971 hCoV-19/England/CAMC-B3609D/2020\|EPI_ISL_645301\|20 | ···················· | ···················· | ···················· | ···················· | ···················· | ···················· | |
| R | 5972 hCoV-19/England/MILK-B3D6EF/2020\|EPI_ISL_645302\|20 | ···················· | ···················· | ···················· | ···················· | ···················· | ···················· | |
| R | 5973 hCoV-19/England/ALDP-B3E45D/2020\|EPI_ISL_645303\|20 | ···················· | ···················· | ···················· | ···················· | ···················· | ···················· | |
| R | 5974 hCoV-19/England/CAMC-B363EF/2020\|EPI_ISL_645304\|20 | ···················· | ···················· | ···················· | ···················· | ···················· | ···················· | |
| R | 5975 hCoV-19/England/CAMC-B370E7/2020\|EPI_ISL_645305\|20 | ···················· | ···················· | ···················· | ···················· | ···················· | ···················· | |
| R | 5976 hCoV-19/England/ALDP-B26397/2020\|EPI_ISL_646621\|20 | ···················· | ·A·················· | ···················· | ···················· | ···················· | ··A················· | |
| R | 5977 hCoV-19/England/ALDP-B2634C/2020\|EPI_ISL_646626\|20 | ···················· | ···················· | ···················· | ···················· | ···················· | ···················· | |
| R | 5978 hCoV-19/England/ALDP-B24FD1/2020\|EPI_ISL_646628\|20 | ···················· | ···················· | ···················· | ···················· | ···················· | ···················· | |
| R | 5979 hCoV-19/England/QEUH-B2B4CC/2020\|EPI_ISL_646630\|20 | ···················· | ···················· | ···················· | ···················· | ·····T·············· | ···················· | |
| R | 5980 hCoV-19/England/ALDP-B24F59/2020\|EPI_ISL_646631\|20 | ···················· | ···················· | ···················· | ···················· | ···················· | ···················· | |
| R | 5981 hCoV-19/England/QEUH-B2B9BE/2020\|EPI_ISL_646632\|20 | ···················· | ···················· | ···················· | ···················· | ···················· | ···················· | |
| R | 5982 hCoV-19/England/QEUH-B2B523/2020\|EPI_ISL_646633\|20 | ···················· | ···················· | ···················· | ···················· | ···················· | ···················· | |
| R | 5983 hCoV-19/England/QEUH-B2B954/2020\|EPI_ISL_646634\|20 | ···················· | ···················· | ···················· | ···················· | ···················· | ···················· | |
| R | 5984 hCoV-19/England/ALDP-B24FE0/2020\|EPI_ISL_646635\|20 | ···················· | ···················· | ···················· | ···················· | ···················· | ···················· | |
| R | 5985 hCoV-19/England/QEUH-B2B471/2020\|EPI_ISL_646636\|20 | ···················· | ···················· | ···················· | ···················· | ···················· | ···················· | |
| R | 5986 hCoV-19/England/ALDP-B2394D/2020\|EPI_ISL_647000\|20 | ···················· | ···················· | ···················· | ···················· | ···················· | ···················· | |
| R | 5987 hCoV-19/England/QEUH-B2D9BC/2020\|EPI_ISL_647001\|20 | ···················· | ···················· | ···················· | ···················· | ···················· | ···················· | |
| R | 5988 hCoV-19/England/ALDP-B24A58/2020\|EPI_ISL_647002\|20 | ···················· | ···················· | ···················· | ···················· | ···················· | ···················· | |
| R | 5989 hCoV-19/England/QEUH-B2D7A3/2020\|EPI_ISL_647003\|20 | ···················· | ···················· | ···················· | ···················· | ···················· | ···················· | |
| R | 5990 hCoV-19/England/ALDP-B249A6/2020\|EPI_ISL_647004\|20 | ···················· | ···················· | ···················· | ···················· | ···················· | ···················· | |
| R | 5991 hCoV-19/England/ALDP-B24B28/2020\|EPI_ISL_647005\|20 | ···················· | ···················· | ···················· | ···················· | ···················· | ···················· | |
| R | 5992 hCoV-19/England/QEUH-B2DB89/2020\|EPI_ISL_647006\|20 | ···················· | ···················· | ···················· | ···················· | ···················· | ···················· | |
| R | 5993 hCoV-19/England/ALDP-B243E4/2020\|EPI_ISL_647007\|20 | ···················· | ···················· | ···················· | ···················· | ···················· | ···················· | |
| R | 5994 hCoV-19/England/ALDP-B238C8/2020\|EPI_ISL_647008\|20 | ···················· | ···················· | ···················· | ···················· | ···················· | ···················· | |
| R | 5995 hCoV-19/England/ALDP-B2419F/2020\|EPI_ISL_647010\|20 | ···················· | ···················· | ···················· | ···················· | ···················· | ···················· | |
| R | 5996 hCoV-19/England/ALDP-B23682/2020\|EPI_ISL_647011\|20 | ···················· | ···················· | ···················· | ···················· | ···················· | ···················· | |
| R | 5997 hCoV-19/England/ALDP-B24478/2020\|EPI_ISL_647012\|20 | ···················· | ···················· | ···················· | ···················· | ···················· | ···················· | |
| R | 5998 hCoV-19/England/QEUH-B2D9DA/2020\|EPI_ISL_647014\|20 | ···················· | ···················· | ···················· | ···················· | ···················· | ···················· | |
| R | 5999 hCoV-19/England/ALDP-B23743/2020\|EPI_ISL_647015\|20 | ···················· | ···················· | ···················· | ···················· | ···················· | ···················· | |
| R | 6000 hCoV-19/England/ALDP-B2452A/2020\|EPI_ISL_647017\|20 | ···················· | ···················· | ···················· | ···················· | ···················· | ···················· | |
| R | 6001 hCoV-19/England/ALDP-B24AA3/2020\|EPI_ISL_647018\|20 | ···················· | ···················· | ···················· | ···················· | ···················· | ···················· | |
| R | 6002 hCoV-19/England/ALDP-B23619/2020\|EPI_ISL_647019\|20 | ···················· | ···················· | ···················· | ···················· | ···················· | ···················· | |
| R | 6003 hCoV-19/England/ALDP-B24C34/2020\|EPI_ISL_647020\|20 | ···················· | ···················· | ···················· | ···················· | ···················· | ···················· | |
| R | 6004 hCoV-19/England/ALDP-B245C0/2020\|EPI_ISL_647021\|20 | ···················· | ·A·················· | ···················· | ···················· | ···················· | ··A················· | |
| R | 6005 hCoV-19/England/ALDP-B24A2B/2020\|EPI_ISL_647022\|20 | ···················· | ···················· | ···················· | ···················· | ···················· | ···················· | |
| R | 6006 hCoV-19/England/QEUH-B2D9CB/2020\|EPI_ISL_647023\|20 | ···················· | ···················· | ···················· | ···················· | ···················· | ···················· | |
| R | 6007 hCoV-19/England/ALDP-B2429C/2020\|EPI_ISL_647024\|20 | ···················· | ···················· | ···················· | ···················· | ···················· | ···················· | |
| R | 6008 hCoV-19/England/ALDP-B23A86/2020\|EPI_ISL_647025\|20 | ···················· | ···················· | ···················· | ···················· | ···················· | ···················· | |
| R | 6009 hCoV-19/England/ALDP-B249C4/2020\|EPI_ISL_647026\|20 | ···················· | ···················· | ···················· | ···················· | ···················· | ···················· | |

| | | | | | | | | |
|---|---|---|---|---|---|---|---|---|
| R | 6010 | hCoV-19/England/ALDP-B242F6/2020\|EPI_ISL_647027\|20 | ···················· | ···················· | ···················· | ···················· | ···················· | ···················· |
| R | 6011 | hCoV-19/England/ALDP-B244D2/2020\|EPI_ISL_647028\|20 | ···················· | ···················· | ···················· | ···················· | ···················· | ···················· |
| R | 6012 | hCoV-19/England/ALDP-B23A3B/2020\|EPI_ISL_647029\|20 | ···················· | ···················· | ···················· | ···················· | ···················· | ···················· |
| R | 6013 | hCoV-19/England/ALDP-B242D8/2020\|EPI_ISL_647030\|20 | ···················· | ···················· | ···················· | ···················· | ···················· | ···················· |
| R | 6014 | hCoV-19/England/QEUH-B2DBB6/2020\|EPI_ISL_647031\|20 | ···················· | ···················· | ···················· | ···················· | ···················· | ···················· |
| R | 6015 | hCoV-19/England/QEUH-B2ED82/2020\|EPI_ISL_647032\|20 | ···················· | ·A·················· | ···················· | ···················· | ···················· | ···················· |
| R | 6016 | hCoV-19/England/ALDP-B24548/2020\|EPI_ISL_647033\|20 | ···················· | ···················· | ···················· | ···················· | ···················· | ···················· |
| R | 6017 | hCoV-19/England/ALDP-B24C8F/2020\|EPI_ISL_647034\|20 | ···················· | ···················· | ···················· | ···················· | ···················· | ···················· |
| R | 6018 | hCoV-19/England/QEUH-B2DB10/2020\|EPI_ISL_647035\|20 | ···················· | ···················· | ···················· | ···················· | ···················· | ···················· |
| R | 6019 | hCoV-19/England/ALDP-B24C25/2020\|EPI_ISL_647037\|20 | ···················· | ···················· | ···················· | ···················· | ···················· | ···················· |
| R | 6020 | hCoV-19/England/ALDP-B236DD/2020\|EPI_ISL_647039\|20 | ···················· | ···················· | ···················· | ···················· | ···················· | ···················· |
| R | 6021 | hCoV-19/England/ALDP-B2385F/2020\|EPI_ISL_647040\|20 | ···················· | ···················· | ···················· | ···················· | ···················· | ···················· |
| R | 6022 | hCoV-19/England/ALDP-B23628/2020\|EPI_ISL_647041\|20 | ···················· | ···················· | ···················· | ···················· | ···················· | ···················· |
| R | 6023 | hCoV-19/England/ALDP-B2486D/2020\|EPI_ISL_647042\|20 | ···················· | ···················· | ···················· | ···················· | ···················· | ···················· |
| R | 6024 | hCoV-19/England/ALDP-B23770/2020\|EPI_ISL_647043\|20 | ···················· | ···················· | ···················· | ···················· | ···················· | ···················· |
| R | 6025 | hCoV-19/England/ALDP-B24575/2020\|EPI_ISL_647044\|20 | ···················· | ···················· | ···················· | ···················· | ···················· | ···················· |
| R | 6026 | hCoV-19/England/QEUH-B2DC2C/2020\|EPI_ISL_647045\|20 | ··T················· | ···················· | ···················· | ···················· | ···················· | ···················· |
| R | 6027 | hCoV-19/England/ALDP-B244B4/2020\|EPI_ISL_647046\|20 | ···················· | ···················· | ···················· | ···················· | ···················· | ···················· |
| R | 6028 | hCoV-19/England/ALDP-B24D13/2020\|EPI_ISL_647047\|20 | ···················· | ···················· | ···················· | ···················· | ···················· | ···················· |
| R | 6029 | hCoV-19/England/ALDP-B24618/2020\|EPI_ISL_647048\|20 | ···················· | ···················· | ···················· | ···················· | ···················· | ···················· |
| R | 6030 | hCoV-19/England/ALDP-B244A5/2020\|EPI_ISL_647049\|20 | ···················· | ···················· | ···················· | ···················· | ···················· | ···················· |
| R | 6031 | hCoV-19/England/QEUH-B2D776/2020\|EPI_ISL_647050\|20 | ···················· | ···················· | ···················· | ···················· | ···················· | ···················· |
| R | 6032 | hCoV-19/England/ALDP-B23BDE/2020\|EPI_ISL_647051\|20 | ···················· | ···················· | ···················· | ···················· | ···················· | ···················· |
| R | 6033 | hCoV-19/England/ALDP-B24584/2020\|EPI_ISL_647052\|20 | ···················· | ···················· | ···················· | ···················· | ···················· | ···················· |
| R | 6034 | hCoV-19/England/ALDP-B239D4/2020\|EPI_ISL_647054\|20 | ···················· | ·A·················· | ···················· | ···················· | ···················· | ···················· |
| R | 6035 | hCoV-19/England/ALDP-B241EA/2020\|EPI_ISL_647056\|20 | ···················· | ···················· | ···················· | ···················· | ···················· | ···················· |
| R | 6036 | hCoV-19/England/ALDP-B24663/2020\|EPI_ISL_647057\|20 | ···················· | ···················· | ···················· | ···················· | ···················· | ···················· |
| R | 6037 | hCoV-19/England/ALDP-B249F1/2020\|EPI_ISL_647058\|20 | ···················· | ···················· | ···················· | ···················· | ···················· | ···················· |
| R | 6038 | hCoV-19/England/QEUH-B2DB7A/2020\|EPI_ISL_647059\|20 | ···················· | ···················· | ···················· | ···················· | ···················· | ···················· |
| R | 6039 | hCoV-19/England/ALDP-B24979/2020\|EPI_ISL_647060\|20 | ···················· | ···················· | ···················· | ···················· | ···················· | ···················· |
| R | 6040 | hCoV-19/England/QEUH-B2DD47/2020\|EPI_ISL_647061\|20 | ···················· | ···················· | ···················· | ···················· | ···················· | ···················· |
| R | 6041 | hCoV-19/England/ALDP-B239A7/2020\|EPI_ISL_647062\|20 | ···················· | ···················· | ···················· | ···················· | ···················· | ···················· |
| R | 6042 | hCoV-19/England/ALDP-B238E6/2020\|EPI_ISL_647063\|20 | ···················· | ···················· | ···················· | ···················· | ···················· | ···················· |
| R | 6043 | hCoV-19/England/ALDP-B2433F/2020\|EPI_ISL_647064\|20 | ···················· | ···················· | ···················· | ···················· | ···················· | ···················· |
| R | 6044 | hCoV-19/England/ALDP-B242E7/2020\|EPI_ISL_647065\|20 | ···················· | ···················· | ···················· | ···················· | ···················· | ···················· |
| R | 6045 | hCoV-19/England/ALDP-B24988/2020\|EPI_ISL_647066\|20 | ···················· | ···················· | ···················· | ···················· | ···················· | ···················· |
| R | 6046 | hCoV-19/England/QEUH-B2DB5C/2020\|EPI_ISL_647067\|20 | ···················· | ···················· | ···················· | ···················· | ···················· | ···················· |
| R | 6047 | hCoV-19/Wales/ALDP-B24BA0/2020\|EPI_ISL_647068\|2020 | ···················· | ···················· | ···················· | ···················· | ···················· | ···················· |
| R | 6048 | hCoV-19/England/ALDP-B247E8/2020\|EPI_ISL_647069\|20 | ···················· | ···················· | ···················· | ···················· | ···················· | ···················· |
| R | 6049 | hCoV-19/England/ALDP-B247AC/2020\|EPI_ISL_647070\|20 | ···················· | ···················· | ···················· | ···················· | ···················· | ···················· |
| R | 6050 | hCoV-19/England/ALDP-B2741B/2020\|EPI_ISL_647072\|20 | ···················· | ···················· | ···················· | ···················· | ···················· | ···················· |
| R | 6051 | hCoV-19/England/ALDP-B24B46/2020\|EPI_ISL_647074\|20 | ···················· | ···················· | ···················· | ···················· | ···················· | ···················· |
| R | 6052 | hCoV-19/England/ALDP-B248E5/2020\|EPI_ISL_647075\|20 | ···················· | ·A·················· | ···················· | ···················· | ···················· | ···················· |
| R | 6053 | hCoV-19/England/QEUH-B2DBE3/2020\|EPI_ISL_647076\|20 | ···················· | ···················· | ···················· | ···················· | ···················· | ···················· |
| R | 6054 | hCoV-19/England/ALDP-B24A0D/2020\|EPI_ISL_647077\|20 | ···················· | ···················· | ···················· | ···················· | ···················· | ···················· |
| R | 6055 | hCoV-19/England/ALDP-B24751/2020\|EPI_ISL_647078\|20 | ···················· | ···················· | ···················· | ···················· | ···················· | ···················· |
| R | 6056 | hCoV-19/England/ALDP-B2389B/2020\|EPI_ISL_647079\|20 | ···················· | ···················· | ···················· | ···················· | ···················· | ···················· |
| R | 6057 | hCoV-19/England/ALDP-B248A9/2020\|EPI_ISL_647080\|20 | ···················· | ···················· | ···················· | ···················· | ···················· | ···················· |
| R | 6058 | hCoV-19/England/ALDP-B238B9/2020\|EPI_ISL_647083\|20 | ···················· | ···················· | ···················· | ···················· | ···················· | ···················· |
| R | 6059 | hCoV-19/England/ALDP-B23761/2020\|EPI_ISL_647084\|20 | ···················· | ···················· | ···················· | ···················· | ···················· | ···················· |
| R | 6060 | hCoV-19/England/ALDP-B2479D/2020\|EPI_ISL_647085\|20 | ···················· | ···················· | ···················· | ···················· | ···················· | ···················· |
| R | 6061 | hCoV-19/England/ALDP-B23734/2020\|EPI_ISL_647086\|20 | ···················· | ···················· | ···················· | ···················· | ···················· | ···················· |
| R | 6062 | hCoV-19/England/QEUH-B2D98F/2020\|EPI_ISL_647087\|20 | ···················· | ···················· | ···················· | ···················· | ···················· | ···················· |
| R | 6063 | hCoV-19/England/ALDP-B24241/2020\|EPI_ISL_647088\|20 | ···················· | ···················· | ···················· | ···················· | ···················· | ···················· |
| R | 6064 | hCoV-19/England/ALDP-B24144/2020\|EPI_ISL_647089\|20 | ···················· | ···················· | ···················· | ···················· | ···················· | ···················· |
| R | 6065 | hCoV-19/England/QEUH-B2DAF5/2020\|EPI_ISL_647090\|20 | ···················· | ···················· | ···················· | ···················· | ···················· | ···················· |
| R | 6066 | hCoV-19/England/ALDP-B24496/2020\|EPI_ISL_647091\|20 | ···················· | ···················· | ···················· | ···················· | ···················· | ···················· |
| R | 6067 | hCoV-19/England/ALDP-B24997/2020\|EPI_ISL_647092\|20 | ···················· | ···················· | ···················· | ···················· | ···················· | ···················· |
| R | 6068 | hCoV-19/England/ALDP-B24557/2020\|EPI_ISL_647093\|20 | ···················· | ···················· | ···················· | ···················· | ···················· | ···················· |
| R | 6069 | hCoV-19/England/ALDP-B24733/2020\|EPI_ISL_647094\|20 | ···················· | ···················· | ···················· | ···················· | ···················· | ···················· |
| R | 6070 | hCoV-19/England/ALDP-B24636/2020\|EPI_ISL_647095\|20 | ···················· | ···················· | ···················· | ···················· | ···················· | ···················· |
| R | 6071 | hCoV-19/England/QEUH-B2DBD4/2020\|EPI_ISL_647097\|20 | ···················· | ···················· | ···················· | ···················· | ···················· | ···················· |
| R | 6072 | hCoV-19/England/ALDP-B24AB2/2020\|EPI_ISL_647098\|20 | ···················· | ···················· | ···················· | ···················· | ···················· | ···················· |
| R | 6073 | hCoV-19/England/ALDP-B23691/2020\|EPI_ISL_647099\|20 | ···················· | ···················· | ···················· | ···················· | ···················· | ···················· |
| R | 6074 | hCoV-19/Wales/QEUH-AD33A9/2020\|EPI_ISL_647950\|2020 | ···················· | ···················· | ···················· | ···················· | ···················· | ···················· |
| R | 6075 | hCoV-19/Wales/QEUH-AD328E/2020\|EPI_ISL_647952\|2020 | ···················· | ···················· | ···················· | ···················· | ···················· | ···················· |
| R | 6076 | hCoV-19/Wales/QEUH-AD09F5/2020\|EPI_ISL_647957\|2020 | ···················· | ···················· | ···················· | ···················· | ···················· | ···················· |
| R | 6077 | hCoV-19/Wales/QEUH-AD3321/2020\|EPI_ISL_647958\|2020 | ···················· | ···················· | ···················· | ···················· | ···················· | ···················· |
| R | 6078 | hCoV-19/Wales/QEUH-AD336D/2020\|EPI_ISL_647959\|2020 | ···················· | ···················· | ···················· | ···················· | ···················· | ···················· |
| R | 6079 | hCoV-19/Wales/QEUH-AD3400/2020\|EPI_ISL_647960\|2020 | ···················· | ···················· | ···················· | ···················· | ···················· | ···················· |
| R | 6080 | hCoV-19/Wales/QEUH-AD4997/2020\|EPI_ISL_647961\|2020 | ···················· | ···················· | ···················· | ···················· | ···················· | ···················· |
| R | 6081 | hCoV-19/Wales/QEUH-AD33E5/2020\|EPI_ISL_647962\|2020 | ···················· | ···················· | ···················· | ···················· | ···················· | ···················· |
| R | 6082 | hCoV-19/Wales/QEUH-AD3075/2020\|EPI_ISL_647963\|2020 | ···················· | ···················· | ···················· | ···················· | ···················· | ···················· |
| R | 6083 | hCoV-19/Wales/QEUH-AD2CDC/2020\|EPI_ISL_647964\|2020 | ···················· | ···················· | ···················· | ···················· | ···················· | ···················· |
| R | 6084 | hCoV-19/Wales/QEUH-AD0764/2020\|EPI_ISL_647965\|2020 | ···················· | ···················· | ···················· | ···················· | ···················· | ···················· |
| R | 6085 | hCoV-19/Wales/QEUH-AD49E2/2020\|EPI_ISL_647966\|2020 | ···················· | ···················· | ···················· | ···················· | ···················· | ···················· |
| R | 6086 | hCoV-19/Wales/QEUH-AD07DD/2020\|EPI_ISL_647967\|2020 | ···················· | ···················· | ···················· | ···················· | ···················· | ···················· |
| R | 6087 | hCoV-19/USA/MD-MDH-0365/2020\|EPI_ISL_648300\|2020-1 | ···················· | ···················· | ···················· | ···················· | ···················· | ···················· |
| R | 6088 | hCoV-19/USA/MD-MDH-0366/2020\|EPI_ISL_648301\|2020-1 | ···················· | ···················· | ···················· | ···················· | ···················· | ···················· |
| R | 6089 | hCoV-19/USA/MD-MDH-0367/2020\|EPI_ISL_648302\|2020-1 | ···················· | ···················· | ···················· | ···················· | ···················· | ···················· |

| | | | | | | | |
|---|---|---|---|---|---|---|---|
| R | 6094 hCoV-19/Equatorial Guinea/12705/2020\|EPI_ISL_64831 | ···················· | ···················· | ···················· | ···················· | ··N················· | ···················· |
| R | 6096 hCoV-19/Equatorial Guinea/2274/2020\|EPI_ISL_648315 | ···················· | ···················· | ···················· | ···················· | ···················· | ···················· |
| R | 6103 hCoV-19/Equatorial Guinea/3350/2020\|EPI_ISL_648326 | ···················· | ···················· | ···················· | ···················· | ···················· | ···················· |
| R | 6112 hCoV-19/Equatorial Guinea/5031/2020\|EPI_ISL_648340 | ···················· | ···················· | ···················· | ···················· | ···················· | ···················· |
| R | 6113 hCoV-19/Equatorial Guinea/5117/2020\|EPI_ISL_648341 | ···················· | ···················· | ···················· | ···················· | ·NN················· | ···················· |
| R | 6115 hCoV-19/Equatorial Guinea/6013/2020\|EPI_ISL_648344 | ···················· | ···················· | ···················· | ···················· | ···················· | ···················· |
| R | 6122 hCoV-19/Equatorial Guinea/9401/2020\|EPI_ISL_648354 | ···················· | ···················· | ···················· | ···················· | ·NN················· | ···················· |
| R | 6128 hCoV-19/USA/CA-CZB-7193/2020\|EPI_ISL_648381\|2020-0 | ···················· | ···················· | ···················· | ···················· | ···················· | ···················· |
| R | 6129 hCoV-19/USA/CA-CZB-7194/2020\|EPI_ISL_648382\|2020-0 | ···················· | ···················· | ···················· | ···················· | ···················· | ···················· |
| R | 6130 hCoV-19/USA/CA-CZB-12289/2020\|EPI_ISL_648384\|2020- | ···················· | ·A·················· | ···················· | ···················· | ···················· | ···················· |
| R | 6131 hCoV-19/USA/CA-CZB-12290/2020\|EPI_ISL_648385\|2020- | ···················· | ···················· | ···················· | ···················· | ···················· | ···················· |
| R | 6132 hCoV-19/USA/CA-CZB-12291/2020\|EPI_ISL_648386\|2020- | ···················· | ···················· | ···················· | ···················· | ···················· | ···················· |
| R | 6133 hCoV-19/USA/CA-CZB-12293/2020\|EPI_ISL_648387\|2020- | ···················· | ···················· | ···················· | ···················· | ··········Y········· | ···················· |
| R | 6134 hCoV-19/USA/CA-CZB-12295/2020\|EPI_ISL_648388\|2020- | ···················· | ···················· | ···················· | ···················· | ···················· | ···················· |
| R | 6135 hCoV-19/USA/CA-CZB-12297/2020\|EPI_ISL_648389\|2020- | ···················· | ·A·················· | ···················· | ···················· | ···················· | ···················· |
| R | 6136 hCoV-19/USA/CA-CZB-12298/2020\|EPI_ISL_648390\|2020- | ···················· | ·A·················· | ···················· | ···················· | ···················· | ···················· |
| R | 6137 hCoV-19/USA/CA-CZB-12299/2020\|EPI_ISL_648391\|2020- | ···················· | ·A·················· | ···················· | ···················· | ···················· | ···················· |
| R | 6138 hCoV-19/USA/CA-CZB-12300/2020\|EPI_ISL_648392\|2020- | ···················· | ·A·················· | ···················· | ···················· | ···················· | ···················· |
| R | 6139 hCoV-19/USA/CA-CZB-12303/2020\|EPI_ISL_648393\|2020- | ···················· | ·A·················· | ···················· | ···················· | ···················· | ···················· |
| R | 6140 hCoV-19/USA/CA-CZB-12305/2020\|EPI_ISL_648394\|2020- | ···················· | ·A·················· | ···················· | ···················· | ···················· | ···················· |
| R | 6141 hCoV-19/USA/CA-CZB-12309/2020\|EPI_ISL_648395\|2020- | ···················· | ·A·················· | ···················· | ···················· | ···················· | ···················· |
| R | 6142 hCoV-19/USA/CA-CZB-12311/2020\|EPI_ISL_648396\|2020- | ···················· | ·A·················· | ···················· | ···················· | ···················· | ···················· |
| R | 6143 hCoV-19/USA/CA-CZB-12313/2020\|EPI_ISL_648397\|2020- | ···················· | ·A·················· | ···················· | ···················· | ···················· | ···················· |
| R | 6145 hCoV-19/England/QEUH-B0DC6E/2020\|EPI_ISL_642000\|20 | ···················· | ···················· | ···················· | ···················· | ···················· | ···················· |
| R | 6146 hCoV-19/England/QEUH-B0E429/2020\|EPI_ISL_642002\|20 | ···················· | ···················· | ···················· | ···················· | ···················· | ···················· |
| R | 6147 hCoV-19/England/QEUH-B0F8B3/2020\|EPI_ISL_642003\|20 | ···················· | ···················· | ···················· | ···················· | ···················· | ···················· |
| R | 6148 hCoV-19/England/QEUH-B1107A/2020\|EPI_ISL_642004\|20 | ···················· | ···················· | ···················· | ···················· | ···················· | ···················· |
| R | 6149 hCoV-19/England/QEUH-B0EB7F/2020\|EPI_ISL_642005\|20 | ···················· | ···················· | ···················· | ···················· | ···················· | ···················· |
| R | 6150 hCoV-19/England/QEUH-B0DD6B/2020\|EPI_ISL_642006\|20 | ···················· | ···················· | ···················· | ···················· | ···················· | ···················· |
| R | 6151 hCoV-19/England/QEUH-B0FC99/2020\|EPI_ISL_642100\|20 | ···················· | ·A·······A·········· | ···················· | ···················· | ···················· | ···················· |
| R | 6152 hCoV-19/England/QEUH-B0EB51/2020\|EPI_ISL_642101\|20 | ···················· | ···················· | ···················· | ···················· | ···················· | ···················· |
| R | 6153 hCoV-19/England/QEUH-B0E465/2020\|EPI_ISL_642102\|20 | ···················· | ···················· | ···················· | ···················· | ···················· | ···················· |
| R | 6154 hCoV-19/England/QEUH-B0E878/2020\|EPI_ISL_642103\|20 | ···················· | ···················· | ···················· | ···················· | ···················· | ···················· |
| R | 6155 hCoV-19/England/QEUH-B0F3C1/2020\|EPI_ISL_642104\|20 | ···················· | ···················· | ···················· | ···················· | ···················· | ···················· |
| R | 6156 hCoV-19/England/QEUH-B0DD2F/2020\|EPI_ISL_642105\|20 | ···················· | ···················· | ···················· | ···················· | ···················· | ···················· |
| R | 6157 hCoV-19/England/QEUH-B0EAEB/2020\|EPI_ISL_642106\|20 | ···················· | ···················· | ···················· | ···················· | ···················· | ···················· |
| R | 6158 hCoV-19/England/QEUH-B0DC40/2020\|EPI_ISL_642107\|20 | ···················· | ·A·················· | ···················· | ···················· | ···················· | ···················· |
| R | 6159 hCoV-19/England/QEUH-B0F938/2020\|EPI_ISL_642108\|20 | ···················· | ···················· | ···················· | ···················· | ···················· | ···················· |
| R | 6160 hCoV-19/England/QEUH-B0E122/2020\|EPI_ISL_642109\|20 | ···················· | ···················· | ···················· | ···················· | ···················· | ···················· |
| R | 6161 hCoV-19/England/QEUH-B0DF56/2020\|EPI_ISL_642110\|20 | ···················· | ···················· | ···················· | ···················· | ···················· | ···················· |
| R | 6162 hCoV-19/England/QEUH-B0FC11/2020\|EPI_ISL_642111\|20 | ···················· | ···················· | ···················· | ···················· | ···················· | ···················· |
| R | 6163 hCoV-19/England/QEUH-B0EC9A/2020\|EPI_ISL_642112\|20 | ···················· | ···················· | ···················· | ···················· | ···················· | ···················· |
| R | 6164 hCoV-19/England/QEUH-B0E69C/2020\|EPI_ISL_642113\|20 | ···················· | ···················· | ···················· | ···················· | ···················· | ···················· |
| R | 6165 hCoV-19/England/QEUH-B0DD4D/2020\|EPI_ISL_642114\|20 | ···················· | ···················· | ···················· | ···················· | ···················· | ···················· |
| R | 6166 hCoV-19/England/QEUH-B0E67E/2020\|EPI_ISL_642115\|20 | ···················· | ···················· | ···················· | ···················· | ···················· | ···················· |
| R | 6167 hCoV-19/England/QEUH-B0FBF6/2020\|EPI_ISL_642133\|20 | ···················· | ···················· | ···················· | ···················· | ···················· | ···················· |
| R | 6168 hCoV-19/England/QEUH-B0E2E3/2020\|EPI_ISL_642138\|20 | ···················· | ···················· | ···················· | ···················· | ····T··············· | ···················· |
| R | 6169 hCoV-19/England/QEUH-B0EABE/2020\|EPI_ISL_642139\|20 | ···················· | ···················· | ···················· | ···················· | ···················· | ···················· |
| R | 6170 hCoV-19/England/QEUH-B0F3EF/2020\|EPI_ISL_642140\|20 | ···················· | ···················· | ···················· | ···················· | ···················· | ···················· |
| R | 6171 hCoV-19/England/QEUH-B0E92A/2020\|EPI_ISL_642141\|20 | ···················· | ···················· | ···················· | ···················· | ···················· | ···················· |
| R | 6172 hCoV-19/England/QEUH-B0DEB3/2020\|EPI_ISL_642142\|20 | ···················· | ···················· | ···················· | ···················· | ···················· | ···················· |
| R | 6173 hCoV-19/England/QEUH-B0F33A/2020\|EPI_ISL_642143\|20 | ···················· | ···················· | ···················· | ···················· | ···················· | ···················· |
| R | 6174 hCoV-19/England/QEUH-B0EBAC/2020\|EPI_ISL_642144\|20 | ···················· | ···················· | ···················· | ···················· | ···················· | ···················· |
| R | 6175 hCoV-19/England/QEUH-B0FCC6/2020\|EPI_ISL_642145\|20 | ···················· | ···················· | ···················· | ···················· | ···················· | ···················· |
| R | 6176 hCoV-19/England/QEUH-B0E447/2020\|EPI_ISL_642146\|20 | ···················· | ···················· | ···················· | ···················· | ···················· | ···················· |
| R | 6177 hCoV-19/England/QEUH-B0DC13/2020\|EPI_ISL_642147\|20 | ···················· | ···················· | ···················· | ···················· | ···················· | ···················· |
| R | 6178 hCoV-19/England/QEUH-B0F886/2020\|EPI_ISL_642148\|20 | ···················· | ·A·················· | ···················· | ···················· | ···················· | ···················· |
| R | 6179 hCoV-19/England/QEUH-B0EBBB/2020\|EPI_ISL_642149\|20 | ···················· | ·A··G··············· | ···················· | ···················· | ···················· | ···················· |
| R | 6180 hCoV-19/England/QEUH-B0DBF8/2020\|EPI_ISL_642150\|20 | ···················· | ·A·················· | ···················· | ···················· | ···················· | ···················· |
| R | 6181 hCoV-19/England/QEUH-B0E30E/2020\|EPI_ISL_642152\|20 | ···················· | ···················· | ···················· | ···················· | ······T············· | ···················· |
| R | 6182 hCoV-19/England/QEUH-B0FC7B/2020\|EPI_ISL_642153\|20 | ···················· | ···················· | ···················· | ···················· | ···················· | ···················· |
| R | 6183 hCoV-19/England/QEUH-B0E5F9/2020\|EPI_ISL_642155\|20 | ···················· | ···················· | ···················· | ···················· | ···················· | ···················· |
| R | 6184 hCoV-19/England/QEUH-B0E3B3/2020\|EPI_ISL_642156\|20 | ···················· | ···················· | ···················· | ···················· | ···················· | ···················· |
| R | 6185 hCoV-19/England/QEUH-B0FC6C/2020\|EPI_ISL_642157\|20 | ···················· | ···················· | ···················· | ···················· | ···················· | ···················· |
| R | 6186 hCoV-19/England/QEUH-B0EA72/2020\|EPI_ISL_642158\|20 | ···················· | ···················· | ···················· | ···················· | ···················· | ···················· |
| R | 6187 hCoV-19/England/QEUH-B0E975/2020\|EPI_ISL_642159\|20 | ···················· | ···················· | ···················· | ···················· | ···················· | ···················· |
| R | 6188 hCoV-19/England/QEUH-B0E298/2020\|EPI_ISL_642160\|20 | ···················· | ·········C·········· | ···················· | ···················· | ···················· | ···················· |
| R | 6189 hCoV-19/England/QEUH-B0F8D1/2020\|EPI_ISL_642161\|20 | ···················· | ···················· | ···················· | ···················· | ···················· | ···················· |
| R | 6190 hCoV-19/England/QEUH-B0E474/2020\|EPI_ISL_642162\|20 | ···················· | ···················· | ···················· | ···················· | ···················· | ···················· |
| R | 6191 hCoV-19/England/QEUH-B0E22F/2020\|EPI_ISL_642163\|20 | ···················· | ···················· | ···················· | ···················· | ···················· | ···················· |
| R | 6192 hCoV-19/England/QEUH-B0DC5F/2020\|EPI_ISL_642164\|20 | ···················· | ···················· | ···················· | ···················· | ···················· | ···················· |
| R | 6193 hCoV-19/England/QEUH-B1104D/2020\|EPI_ISL_642165\|20 | ···················· | ···················· | ···················· | ···················· | ···················· | ···················· |
| R | 6194 hCoV-19/England/QEUH-B0DF47/2020\|EPI_ISL_642166\|20 | ···················· | ···················· | ···················· | ···················· | ···················· | ···················· |
| R | 6195 hCoV-19/England/QEUH-B0DADD/2020\|EPI_ISL_642167\|20 | ···················· | ···················· | ···················· | ···················· | ···················· | ···················· |
| R | 6196 hCoV-19/England/QEUH-B0DB52/2020\|EPI_ISL_642168\|20 | ···················· | ·A·················· | ···················· | ···················· | ···················· | ···················· |
| R | 6197 hCoV-19/England/QEUH-B0E5DB/2020\|EPI_ISL_642169\|20 | ···················· | ···················· | ···················· | ···················· | ···················· | ···················· |
| R | 6198 hCoV-19/England/QEUH-B0DF65/2020\|EPI_ISL_642170\|20 | ···················· | ···················· | ···················· | ···················· | ···················· | ···················· |
| R | 6199 hCoV-19/England/QEUH-B0E553/2020\|EPI_ISL_642171\|20 | ···················· | ···················· | ···················· | ···················· | ···················· | ···················· |
| R | 6200 hCoV-19/England/QEUH-B0DA19/2020\|EPI_ISL_642173\|20 | ···················· | ···················· | ···················· | ···················· | ···················· | ···················· |
| R | 6201 hCoV-19/England/QEUH-B0E869/2020\|EPI_ISL_642174\|20 | ···················· | ···················· | ···················· | ···················· | ···················· | ···················· |

| | | | | | | | |
|---|---|---|---|---|---|---|---|
| R | 6202 hCoV-19/England/QEUH-B0E1C8/2020\|EPI_ISL_642175\|20 | ···················· | ···················· | ···················· | ···················· | ···················· | ···················· |
| R | 6203 hCoV-19/England/QEUH-B0DD3E/2020\|EPI_ISL_642176\|20 | ···················· | ···················· | ···················· | ···················· | ···················· | ···················· |
| R | 6204 hCoV-19/England/QEUH-B0F90B/2020\|EPI_ISL_642177\|20 | ···················· | ·A·················· | ···················· | ···················· | ···················· | ···················· |
| R | 6205 hCoV-19/England/QEUH-B0DF0B/2020\|EPI_ISL_642178\|20 | ···················· | ···················· | ···················· | ···················· | ···················· | ···················· |
| R | 6206 hCoV-19/England/QEUH-B11089/2020\|EPI_ISL_642179\|20 | ···················· | ···················· | ···················· | ···················· | ···················· | ···················· |
| R | 6207 hCoV-19/England/QEUH-B0FC3F/2020\|EPI_ISL_642180\|20 | ···················· | ···················· | ···················· | ···················· | ···················· | ···················· |
| R | 6208 hCoV-19/England/QEUH-B0E517/2020\|EPI_ISL_642181\|20 | ···················· | ···················· | ···················· | ···················· | ···················· | ···················· |
| R | 6209 hCoV-19/England/QEUH-B0FCB7/2020\|EPI_ISL_642182\|20 | ···················· | ···················· | ···················· | ···················· | ···················· | ···················· |
| R | 6210 hCoV-19/England/QEUH-B0F929/2020\|EPI_ISL_642183\|20 | ···················· | ···················· | ···················· | ···················· | ···················· | ···················· |
| R | 6211 hCoV-19/England/QEUH-B0E9FD/2020\|EPI_ISL_642184\|20 | ···················· | ···················· | ···················· | ···················· | ···················· | ···················· |
| R | 6212 hCoV-19/England/QEUH-B0DFFC/2020\|EPI_ISL_642185\|20 | ···················· | ···················· | ···················· | ···················· | ···················· | ···················· |
| R | 6213 hCoV-19/England/QEUH-B0EAFA/2020\|EPI_ISL_642186\|20 | ···················· | ···················· | ···················· | ···················· | ···················· | ···················· |
| R | 6214 hCoV-19/England/QEUH-B0E3A4/2020\|EPI_ISL_642187\|20 | ···················· | ···················· | ···················· | ···················· | ···················· | ···················· |
| R | 6215 hCoV-19/England/QEUH-B0DF74/2020\|EPI_ISL_642188\|20 | ···················· | ···················· | ···················· | ···················· | ···················· | ···················· |
| R | 6216 hCoV-19/England/QEUH-B0E377/2020\|EPI_ISL_642189\|20 | ···················· | ···················· | ···················· | ···················· | ···················· | ···················· |
| R | 6217 hCoV-19/England/QEUH-B0E368/2020\|EPI_ISL_642190\|20 | ···················· | ···················· | ···················· | ···················· | ···················· | ···················· |
| R | 6218 hCoV-19/England/QEUH-B0DFB0/2020\|EPI_ISL_642191\|20 | ···················· | ···················· | ···················· | ···················· | ···················· | ···················· |
| R | 6219 hCoV-19/England/QEUH-B0E641/2020\|EPI_ISL_642192\|20 | ···················· | ···················· | ···················· | ···················· | ···················· | ···················· |
| R | 6220 hCoV-19/England/QEUH-B0ECA9/2020\|EPI_ISL_642193\|20 | ···················· | ···················· | ···················· | ···················· | ···················· | ···················· |
| R | 6221 hCoV-19/England/QEUH-B0DBBC/2020\|EPI_ISL_642195\|20 | ···················· | ···················· | ···················· | ···················· | ···················· | ···················· |
| R | 6222 hCoV-19/England/QEUH-B0E984/2020\|EPI_ISL_642196\|20 | ···················· | ···················· | ···················· | ···················· | ···················· | ···················· |
| R | 6223 hCoV-19/England/QEUH-B0EB9D/2020\|EPI_ISL_642197\|20 | ···················· | ···················· | ···················· | ···················· | ···················· | ···················· |
| R | 6224 hCoV-19/England/QEUH-B0F8C2/2020\|EPI_ISL_642198\|20 | ···················· | ···················· | ···················· | ···················· | ···················· | ···················· |
| R | 6225 hCoV-19/England/QEUH-B0E395/2020\|EPI_ISL_642199\|20 | ···················· | ···················· | ···················· | ···················· | ····T··············· | ···················· |
| R | 6226 hCoV-19/England/CAMC-B0CE1E/2020\|EPI_ISL_643326\|20 | ···················· | ···················· | ···················· | ···················· | ···················· | ···················· |
| R | 6227 hCoV-19/England/CAMC-B0C449/2020\|EPI_ISL_643327\|20 | ···················· | ···················· | ···················· | ···················· | ···················· | ···················· |
| R | 6228 hCoV-19/England/CAMC-B0AFD1/2020\|EPI_ISL_643329\|20 | ···················· | ···················· | ···················· | ···················· | ······T············· | ···················· |
| R | 6229 hCoV-19/England/CAMC-B0B6EA/2020\|EPI_ISL_643330\|20 | ···················· | ···················· | ···················· | ···················· | ···················· | ···················· |
| R | 6230 hCoV-19/England/CAMC-B0AEF2/2020\|EPI_ISL_643331\|20 | ···················· | ·A·················· | ···················· | ···················· | ···················· | ···················· |
| R | 6231 hCoV-19/England/CAMC-B0D062/2020\|EPI_ISL_643332\|20 | ···················· | ·A·················· | ···················· | ···················· | ···················· | ···A················ |
| R | 6232 hCoV-19/England/CAMC-B0ADD7/2020\|EPI_ISL_643333\|20 | ···················· | ···················· | ···················· | ···················· | ······T············· | ···················· |
| R | 6233 hCoV-19/England/CAMC-B07B0D/2020\|EPI_ISL_643334\|20 | ···················· | ···················· | ···················· | ···················· | ···················· | ···················· |
| R | 6234 hCoV-19/England/CAMC-B0AC34/2020\|EPI_ISL_643335\|20 | ···················· | ···················· | ···················· | ···················· | ······T············· | ···················· |
| R | 6235 hCoV-19/England/CAMC-B0CA92/2020\|EPI_ISL_643336\|20 | ···················· | ···················· | ···················· | ···················· | ···················· | ···················· |
| R | 6236 hCoV-19/England/CAMC-B0C68F/2020\|EPI_ISL_643337\|20 | ···················· | ···················· | ···················· | ···················· | ···················· | ···················· |
| R | 6237 hCoV-19/England/CAMC-B0C801/2020\|EPI_ISL_643400\|20 | ···················· | ·A·················· | ···················· | ···················· | ···················· | ···················· |
| R | 6238 hCoV-19/England/CAMC-B0C8D4/2020\|EPI_ISL_643402\|20 | ···················· | ···················· | ···················· | ···················· | ···················· | ···················· |
| R | 6239 hCoV-19/England/CAMC-B080FE/2020\|EPI_ISL_643403\|20 | ···················· | ···················· | ···················· | ···················· | ···················· | ···················· |
| R | 6240 hCoV-19/England/CAMC-B0CD7B/2020\|EPI_ISL_643404\|20 | ···················· | ···················· | ···················· | ···················· | ···················· | ···················· |
| R | 6241 hCoV-19/England/CAMC-B0AF0E/2020\|EPI_ISL_643405\|20 | ···················· | ···················· | ···················· | ···················· | ···················· | ···················· |
| R | 6242 hCoV-19/England/CAMC-B0CC32/2020\|EPI_ISL_643406\|20 | ···················· | ···················· | ···················· | ···················· | ···················· | ···················· |
| R | 6243 hCoV-19/England/CAMC-B0B107/2020\|EPI_ISL_643408\|20 | ···················· | ···················· | ···················· | ···················· | ···················· | ···················· |
| R | 6244 hCoV-19/England/CAMC-B0AF2C/2020\|EPI_ISL_643409\|20 | ···················· | ···················· | ···················· | ···················· | ···················· | ···················· |
| R | 6245 hCoV-19/England/CAMC-B08B39/2020\|EPI_ISL_643410\|20 | ···················· | ···················· | ···················· | ···················· | ···················· | ···················· |
| R | 6246 hCoV-19/England/CAMC-B0CC6F/2020\|EPI_ISL_643411\|20 | ···················· | ···················· | ···················· | ···················· | ···················· | ···················· |
| R | 6247 hCoV-19/England/CAMC-B07DDA/2020\|EPI_ISL_643412\|20 | ···················· | ···················· | ···················· | ···················· | ···················· | ···················· |
| R | 6248 hCoV-19/England/CAMC-B0B310/2020\|EPI_ISL_643413\|20 | ···················· | ···················· | ···················· | ···················· | ···················· | ···················· |
| R | 6249 hCoV-19/England/CAMC-B0B28C/2020\|EPI_ISL_643414\|20 | ···················· | ···················· | ···················· | ···················· | ···················· | ···················· |
| R | 6250 hCoV-19/England/CAMC-B0A89A/2020\|EPI_ISL_643415\|20 | ···················· | ···················· | ···················· | ···················· | ······T············· | ···················· |
| R | 6251 hCoV-19/England/CAMC-B07E31/2020\|EPI_ISL_643416\|20 | ···················· | ···················· | ···················· | ···················· | ···················· | ···················· |
| R | 6252 hCoV-19/England/CAMC-B0B037/2020\|EPI_ISL_643417\|20 | ···················· | ···················· | ···················· | ···················· | ···················· | ···················· |
| R | 6253 hCoV-19/England/CAMC-B0B046/2020\|EPI_ISL_643418\|20 | ···················· | ···················· | ···················· | ···················· | ···················· | ···················· |
| R | 6254 hCoV-19/England/CAMC-B0B750/2020\|EPI_ISL_643419\|20 | ···················· | ···················· | ···················· | ···················· | ···················· | ···················· |
| R | 6255 hCoV-19/England/CAMC-B08559/2020\|EPI_ISL_643420\|20 | ···················· | ···················· | ···················· | ···················· | ···················· | ···················· |
| R | 6256 hCoV-19/England/CAMC-B0AE01/2020\|EPI_ISL_643422\|20 | ···················· | ···················· | ···················· | ···················· | ···················· | ···················· |
| R | 6257 hCoV-19/England/CAMC-B0B4D1/2020\|EPI_ISL_643423\|20 | ···················· | ···················· | ···················· | ···················· | ···················· | ···················· |
| R | 6258 hCoV-19/England/CAMC-B0AFE0/2020\|EPI_ISL_643424\|20 | ···················· | ···················· | ···················· | ···················· | ···················· | ···················· |
| R | 6259 hCoV-19/England/CAMC-B080C1/2020\|EPI_ISL_643425\|20 | ···················· | ···················· | ···················· | ···················· | ···················· | ···················· |
| R | 6260 hCoV-19/England/CAMC-B0CC14/2020\|EPI_ISL_643426\|20 | ···················· | ···················· | ···················· | ···················· | ······T············· | ···················· |
| R | 6261 hCoV-19/England/CAMC-B0839B/2020\|EPI_ISL_643427\|20 | ···················· | ·A·················· | ···················· | ···················· | ···················· | ···A················ |
| R | 6262 hCoV-19/England/CAMC-B07B2B/2020\|EPI_ISL_643428\|20 | ···················· | ···················· | ···················· | ···················· | ···················· | ···················· |
| R | 6263 hCoV-19/England/CAMC-B0B34D/2020\|EPI_ISL_643429\|20 | ···················· | ···················· | ···················· | ···················· | ···················· | ···················· |
| R | 6264 hCoV-19/England/CAMC-B07DF8/2020\|EPI_ISL_643430\|20 | ···················· | ···················· | ···················· | ···················· | ···················· | ···················· |
| R | 6265 hCoV-19/England/CAMC-B0AE5C/2020\|EPI_ISL_643431\|20 | ···················· | ···················· | ···················· | ···················· | ···················· | ···················· |
| R | 6266 hCoV-19/England/CAMC-B0B32F/2020\|EPI_ISL_643432\|20 | ···················· | ···················· | ···················· | ···················· | ···················· | ···················· |
| R | 6267 hCoV-19/England/CAMC-B0CBBD/2020\|EPI_ISL_643433\|20 | ···················· | ···················· | ···················· | ···················· | ···················· | ···················· |
| R | 6268 hCoV-19/England/CAMC-B0C4B2/2020\|EPI_ISL_643434\|20 | ···················· | ···················· | ···················· | ···················· | ···················· | ···················· |
| R | 6269 hCoV-19/England/CAMC-B0B1BC/2020\|EPI_ISL_643435\|20 | ···················· | ···················· | ···················· | ···················· | ···················· | ···················· |
| R | 6270 hCoV-19/England/CAMC-B0B723/2020\|EPI_ISL_643436\|20 | ···················· | ···················· | ···················· | ···················· | ······T············· | ···················· |
| R | 6271 hCoV-19/England/CAMC-B0CB53/2020\|EPI_ISL_643439\|20 | ···················· | ···················· | ···················· | ···················· | ···················· | ···················· |
| R | 6272 hCoV-19/England/CAMC-B0CCC9/2020\|EPI_ISL_643440\|20 | ···················· | ···················· | ···················· | ···················· | ···················· | ···················· |
| R | 6273 hCoV-19/England/CAMC-B0B5DE/2020\|EPI_ISL_643441\|20 | ···················· | ·A·················· | ···················· | ···················· | ···················· | ···A················ |
| R | 6274 hCoV-19/England/CAMC-B0AFC2/2020\|EPI_ISL_643442\|20 | ···················· | ···················· | ···················· | ···················· | ···················· | ···················· |
| R | 6275 hCoV-19/England/CAMC-B0C8A7/2020\|EPI_ISL_643443\|20 | ···················· | ···················· | ···················· | ···················· | ···················· | ···················· |
| R | 6276 hCoV-19/England/CAMC-B0895D/2020\|EPI_ISL_643444\|20 | ···················· | ···················· | ···················· | ···················· | ···················· | ···················· |
| R | 6277 hCoV-19/England/CAMC-B0CB62/2020\|EPI_ISL_643445\|20 | ···················· | ···················· | ···················· | ···················· | ···················· | ···················· |
| R | 6278 hCoV-19/England/CAMC-B0B019/2020\|EPI_ISL_643446\|20 | ···················· | ···················· | ···················· | ···················· | ······T············· | ···················· |
| R | 6279 hCoV-19/England/CAMC-B0CC41/2020\|EPI_ISL_643469\|20 | ···················· | ···················· | ···················· | ···················· | ···················· | ···················· |
| R | 6280 hCoV-19/England/CAMC-B0B27D/2020\|EPI_ISL_643470\|20 | ···················· | ···················· | ···················· | ···················· | ···················· | ···················· |
| R | 6281 hCoV-19/England/CAMC-B0B2F5/2020\|EPI_ISL_643471\|20 | ···················· | ···················· | ···················· | ···················· | ···················· | ···················· |

R 6282 hCoV-19/England/CAMC-B0AC25/2020|EPI_ISL_643473|20 ···················· ···················· ····················· ···················· ···················· ····················
R 6283 hCoV-19/England/CAMC-B0BA84/2020|EPI_ISL_643474|20 ···················· ···················· ····················· ···················· ······T············· ····················
R 6284 hCoV-19/England/CAMC-B08085/2020|EPI_ISL_643476|20 ···················· ···················· ····················· ···················· ···················· ····················
R 6285 hCoV-19/England/CAMC-B0AAEF/2020|EPI_ISL_643477|20 ···················· ···················· ····················· ···················· ···················· ····················
R 6286 hCoV-19/England/CAMC-B07E8C/2020|EPI_ISL_643478|20 ···················· ···················· ····················· ···················· ···················· ····················
R 6287 hCoV-19/England/CAMC-B08261/2020|EPI_ISL_643479|20 ···················· ···················· ····················· ···················· ······T············· ····················
R 6288 hCoV-19/England/CAMC-B0B25F/2020|EPI_ISL_643480|20 ···················· ···················· ····················· ···················· ···················· ····················
R 6289 hCoV-19/England/CAMC-B07B3A/2020|EPI_ISL_643481|20 ···················· ···················· ····················· ···················· ···················· ····················
R 6290 hCoV-19/England/CAMC-B07DAD/2020|EPI_ISL_643482|20 ···················· ···················· ····················· ···················· ······T············· ····················
R 6291 hCoV-19/England/CAMC-B0B41D/2020|EPI_ISL_643483|20 ···················· ·A·················· ····················· ···················· ···················· ···A················
R 6292 hCoV-19/England/CAMC-B0A8E5/2020|EPI_ISL_643484|20 ···················· ···················· ····················· ···················· ···················· ····················
R 6293 hCoV-19/England/CAMC-B0AFB3/2020|EPI_ISL_643485|20 ···················· ···················· ····················· ···················· ···················· ····················
R 6294 hCoV-19/England/CAMC-B0CB44/2020|EPI_ISL_643486|20 ···················· ···················· ····················· ···················· ···················· ····················
R 6295 hCoV-19/England/CAMC-B0B4FF/2020|EPI_ISL_643488|20 ···················· ···················· ····················· ···················· ···················· ····················
R 6296 hCoV-19/England/CAMC-B08128/2020|EPI_ISL_643489|20 ···················· ···················· ····················· ···················· ···················· ····················
R 6297 hCoV-19/England/CAMC-B0AD40/2020|EPI_ISL_643490|20 ···················· ···················· ····················· ···················· ···················· ····················
R 6298 hCoV-19/England/CAMC-B0810A/2020|EPI_ISL_643491|20 ···················· ···················· ····················· ···················· ······T············· ····················
R 6299 hCoV-19/England/CAMC-B0AED4/2020|EPI_ISL_643492|20 ···················· ···················· ····················· ···················· ···················· ····················
R 6300 hCoV-19/England/CAMC-B0CD20/2020|EPI_ISL_643493|20 ···················· ···················· ····················· ···················· ···················· ····················
R 6301 hCoV-19/England/CAMC-B0B0FB/2020|EPI_ISL_643494|20 ···················· ···················· ····················· ···················· ···················· ····················
R 6302 hCoV-19/England/CAMC-B0B459/2020|EPI_ISL_643495|20 ···················· ···················· ····················· ···················· ···················· ····················
R 6303 hCoV-19/England/CAMC-B0AD6E/2020|EPI_ISL_643496|20 ···················· ···················· ····················· ···················· ···················· ····················
R 6304 hCoV-19/England/CAMC-B0AD8C/2020|EPI_ISL_643497|20 ···················· ···················· ····················· ···················· ···················· ····················
R 6305 hCoV-19/England/CAMC-B0AAD0/2020|EPI_ISL_643498|20 ···················· ···················· ····················· ···················· ···················· ····················
R 6306 hCoV-19/England/CAMC-B0B76F/2020|EPI_ISL_643499|20 ···················· ···················· ····················· ···················· ···················· ····················
R 6307 hCoV-19/France/OCC-SC531/2020|EPI_ISL_644700|2020- ···················· ···················· ····················· ···················· ···················· ····················
R 6308 hCoV-19/France/OCC-SC532/2020|EPI_ISL_644701|2020- ···················· ···················· ····················· ···················· ···················· ····················
R 6309 hCoV-19/France/OCC-SC533/2020|EPI_ISL_644702|2020- ···················· ···················· ····················· ···················· ···················· ····················
R 6310 hCoV-19/France/PAC-SC435/2020|EPI_ISL_644703|2020- ···················· ···················· ····················· ···················· ···················· ····················
R 6311 hCoV-19/France/PAC-SC436/2020|EPI_ISL_644704|2020- ···················· ···················· ····················· ···················· ···················· ····················
R 6312 hCoV-19/France/PAC-SC437/2020|EPI_ISL_644705|2020- ···················· ···················· ····················· ···················· ···················· ····················
R 6313 hCoV-19/France/PAC-SC438/2020|EPI_ISL_644706|2020- ···················· ···················· ····················· ···················· ···················· ····················
R 6314 hCoV-19/France/PAC-SC442/2020|EPI_ISL_644707|2020- ···················· ···················· ····················· ···················· ···················· ····················
R 6315 hCoV-19/France/PAC-SC443/2020|EPI_ISL_644708|2020- ···················· ···················· ····················· ···················· ···················· ····················
R 6316 hCoV-19/France/PAC-SC445/2020|EPI_ISL_644709|2020- ···················· ···················· ····················· ···················· ···················· ····················
R 6317 hCoV-19/France/PAC-SC446/2020|EPI_ISL_644710|2020- ···················· ···················· ····················· ···················· ···················· ····················
R 6318 hCoV-19/France/PAC-SC448/2020|EPI_ISL_644711|2020- ···················· ···················· ····················· ···················· ···················· ····················
R 6319 hCoV-19/France/PAC-SC449/2020|EPI_ISL_644712|2020- ···················· ···················· ····················· ···················· ···················· ····················
R 6320 hCoV-19/France/PAC-SC451/2020|EPI_ISL_644713|2020- ···················· ···················· ····················· ···················· ···················· ····················
R 6321 hCoV-19/France/PAC-SC452/2020|EPI_ISL_644714|2020- ···················· ···················· ····················· ···················· ···················· ····················
R 6322 hCoV-19/India/TG-CCMB-OM21/2020|EPI_ISL_644715|202 ···················· ·A·················· ····················· ···················· ···················· ···A················
R 6323 hCoV-19/India/TG-CCMB-OM22/2020|EPI_ISL_644716|202 ···················· ·A·················· ····················· ···················· ···················· ···A················
R 6324 hCoV-19/India/TG-CCMB-OM23/2020|EPI_ISL_644717|202 ···················· ···················· ····················· ···················· ···················· ····················
R 6325 hCoV-19/India/TG-CCMB-OM24/2020|EPI_ISL_644718|202 ···················· ···················· ····················· ···················· ···················· ····················
R 6326 hCoV-19/India/TG-CCMB-OM25/2020|EPI_ISL_644719|202 ···················· ···················· ····················· ···················· ···················· ····················
R 6327 hCoV-19/India/TG-CCMB-OM26/2020|EPI_ISL_644720|202 ······G············· ···················· ····················· ···················· ···················· ····················
R 6328 hCoV-19/India/TG-CCMB-OM27/2020|EPI_ISL_644721|202 ···················· ·A·················· ····················· ···················· ···················· ···A················
R 6329 hCoV-19/India/TG-CCMB-OM28/2020|EPI_ISL_644722|202 ···················· ···················· ····················· ···················· ···················· ····················
R 6330 hCoV-19/India/TG-CCMB-OM29/2020|EPI_ISL_644723|202 ···················· ·A·················· ····················· ···················· ···················· ···A················
R 6331 hCoV-19/India/TG-CCMB-OM30/2020|EPI_ISL_644724|202 ···················· ···················· ····················· ···················· ···················· ····················
R 6332 hCoV-19/India/TG-CCMB-OM31/2020|EPI_ISL_644725|202 ···················· ···················· ····················· ···················· ···················· ····················
R 6333 hCoV-19/India/TG-CCMB-OM32/2020|EPI_ISL_644726|202 ···················· ···················· ····················· ···················· ···················· ····················
R 6334 hCoV-19/India/TG-CCMB-OM33/2020|EPI_ISL_644727|202 ···················· ···················· ····················· ···················· ···················· ····················
R 6335 hCoV-19/India/TG-CCMB-OM34/2020|EPI_ISL_644728|202 ···················· ···················· ····················· ···················· ···················· ····················
R 6336 hCoV-19/India/TG-CCMB-OM35/2020|EPI_ISL_644729|202 ···················· ···················· ····················· ···················· ···················· ····················
R 6337 hCoV-19/India/TG-CCMB-OM36/2020|EPI_ISL_644730|202 ···················· ···················· ····················· ···················· ···················· ····················
R 6338 hCoV-19/India/TG-CCMB-OM37/2020|EPI_ISL_644731|202 ···················· ···················· ····················· ···················· ···················· ····················
R 6339 hCoV-19/India/TG-CCMB-OM38/2020|EPI_ISL_644732|202 ···················· ···················· ····················· ···················· ···················· ····················
R 6340 hCoV-19/India/TG-CCMB-OM39/2020|EPI_ISL_644733|202 ···················· ···················· ····················· ···················· ···················· ····················
R 6341 hCoV-19/India/TG-CCMB-OM40/2020|EPI_ISL_644734|202 ···················· ···················· ····················· ···················· ···················· ····················
R 6342 hCoV-19/England/ALDP-B1F7FF/2020|EPI_ISL_645988|20 ···················· ···················· ····················· ···················· ···················· ····················
R 6343 hCoV-19/England/CAMC-B21C73/2020|EPI_ISL_645989|20 ···················· ···················· ····················· ···················· ···················· ····················
R 6344 hCoV-19/England/ALDP-B1AEA4/2020|EPI_ISL_645990|20 ···················· ···················· ····················· ···················· ···················· ····················
R 6345 hCoV-19/Wales/ALDP-B1FA6F/2020|EPI_ISL_645991|2020 ···················· ···················· ····················· ···················· ···················· ····················
R 6346 hCoV-19/England/CAMC-B22137/2020|EPI_ISL_645992|20 ···················· ···················· ····················· ···················· ······T············· ····················
R 6347 hCoV-19/England/CAMC-B221FB/2020|EPI_ISL_645993|20 ···················· ···················· ····················· ···················· ···················· ····················
R 6348 hCoV-19/England/CAMC-B22225/2020|EPI_ISL_645994|20 ···················· ·A·················· ····················· ···················· ···················· ···A················
R 6349 hCoV-19/England/CAMC-B21E8C/2020|EPI_ISL_645995|20 ···················· ···················· ····················· ···················· ···················· ····················
R 6350 hCoV-19/England/ALDP-B1C0F7/2020|EPI_ISL_645996|20 ···················· ···················· ····················· ···················· ···················· ····················
R 6351 hCoV-19/England/ALDP-B1A3B4/2020|EPI_ISL_645997|20 ···················· ···················· ····················· ···················· ···················· ····················
R 6352 hCoV-19/England/CAMC-B22340/2020|EPI_ISL_645998|20 ···················· ···················· ····················· ···················· ······T············· ····················
R 6353 hCoV-19/England/ALDP-B1DB8C/2020|EPI_ISL_645999|20 ···················· ···················· ····················· ···················· ···················· ····················
R 6354 hCoV-19/Sweden/20-53238/2020|EPI_ISL_648200|2020-1 ···················· ···················· ····················· ···················· ······T············· ····················
R 6355 hCoV-19/Sweden/20-53242/2020|EPI_ISL_648201|2020-1 ···················· ···················· ····················· ···················· ······T············· ····················
R 6356 hCoV-19/Sweden/20-53241/2020|EPI_ISL_648202|2020-1 ···················· ···················· ····················· ···················· ······T············· ····················
R 6357 hCoV-19/Sweden/20-53239/2020|EPI_ISL_648203|2020-1 ···················· ···············G···· ····················· ···················· ···················· ····················
R 6358 hCoV-19/Sweden/20-52711/2020|EPI_ISL_648204|2020-0 ···················· ···················· ····················· ···················· ···················· ····················
R 6359 hCoV-19/Sweden/20-53005/2020|EPI_ISL_648205|2020-0 ···················· ···················· ····················· ···················· ···················· ····················
R 6360 hCoV-19/Sweden/20-53009/2020|EPI_ISL_648206|2020-0 ···················· ···················· ····················· ···················· ···················· ····················
R 6361 hCoV-19/Sweden/20-53232/2020|EPI_ISL_648207|2020-1 ···················· ···················· ····················· ···················· ···················· ····················

| | | | | | | | |
|---|---|---|---|---|---|---|---|
| R | 6362 hCoV-19/Sweden/20-53233/2020\|EPI_ISL_648208\|2020-1 | ···················· | ···················· | ···················· | ···················· | ···················· | ···················· |
| R | 6363 hCoV-19/Argentina/argenTAG-12C0A026/2020\|EPI_ISL_6 | ···················· | ···················· | ···················· | ···················· | ···················· | ···················· |
| R | 6364 hCoV-19/Argentina/argenTAG-12BBC055/2020\|EPI_ISL_6 | ···················· | ···················· | ···················· | ···················· | ···················· | ···················· |
| R | 6365 hCoV-19/Argentina/argenTAG-1283F02A/2020\|EPI_ISL_6 | ···················· | ···················· | ···················· | ···················· | ···················· | ···················· |
| R | 6366 hCoV-19/Argentina/argenTAG-12568054/2020\|EPI_ISL_6 | ···················· | ···················· | ···················· | ···················· | ···················· | ···················· |
| R | 6367 hCoV-19/Argentina/argenTAG-12E7E043/2020\|EPI_ISL_6 | ···················· | ···················· | ···················· | ···················· | ···················· | ···················· |
| R | 6368 hCoV-19/Argentina/argenTAG-12926008/2020\|EPI_ISL_6 | ···················· | ···················· | ···················· | ···················· | ···················· | ···················· |
| R | 6369 hCoV-19/Argentina/argenTAG-126BE00A/2020\|EPI_ISL_6 | ···················· | ···················· | ···················· | ···················· | ···················· | ···················· |
| R | 6370 hCoV-19/Argentina/argenTAG-12CB103F/2020\|EPI_ISL_6 | ···················· | ···················· | ···················· | ···················· | ···················· | ···················· |
| R | 6371 hCoV-19/USA/MA-JLL-865452S/2020\|EPI_ISL_648265\|202 | ···················· | ···················· | ···················· | ···················· | ···················· | ···················· |
| R | 6372 hCoV-19/USA/MA-JLL-383468S/2020\|EPI_ISL_648267\|202 | ···················· | ···················· | ···················· | ···················· | ···················· | ···················· |
| R | 6373 hCoV-19/USA/MA-JLL-796350S/2020\|EPI_ISL_648270\|202 | ···················· | ···················· | ···················· | ···················· | ···················· | ···················· |
| R | 6374 hCoV-19/USA/MD-MDH-0325/2020\|EPI_ISL_648271\|2020-1 | ···················· | ···················· | ···················· | ···················· | ···················· | ···················· |
| R | 6375 hCoV-19/USA/MD-MDH-0327/2020\|EPI_ISL_648272\|2020-1 | ···················· | ·A·················· | ···················· | ···················· | ···················· | ···················· |
| R | 6376 hCoV-19/USA/MD-MDH-0328/2020\|EPI_ISL_648273\|2020-1 | ···················· | ···················· | ···················· | ···················· | ···················· | ···················· |
| R | 6377 hCoV-19/USA/MD-MDH-0329/2020\|EPI_ISL_648274\|2020-1 | ···················· | ···················· | ···················· | ···················· | ···················· | ···················· |
| R | 6378 hCoV-19/USA/MD-MDH-0330/2020\|EPI_ISL_648275\|2020-1 | ···················· | ···················· | ···················· | ···················· | ···················· | ···················· |
| R | 6379 hCoV-19/USA/MD-MDH-0331/2020\|EPI_ISL_648276\|2020-1 | ···················· | ·A·················· | ···················· | ···················· | ···················· | ···················· |
| R | 6380 hCoV-19/USA/MD-MDH-0332/2020\|EPI_ISL_648277\|2020-1 | ···················· | ···················· | ···················· | ···················· | ···················· | ···················· |
| R | 6381 hCoV-19/USA/MD-MDH-0333/2020\|EPI_ISL_648278\|2020-1 | ···················· | ···················· | ···················· | ···················· | ···················· | ···················· |
| R | 6382 hCoV-19/USA/MD-MDH-0334/2020\|EPI_ISL_648279\|2020-1 | ···················· | ···················· | ···················· | ···················· | ···················· | ···················· |
| R | 6383 hCoV-19/USA/MD-MDH-0336/2020\|EPI_ISL_648280\|2020-1 | ···················· | ···········C········ | ···················· | ···················· | ···················· | ···················· |
| R | 6384 hCoV-19/USA/MD-MDH-0338/2020\|EPI_ISL_648281\|2020-1 | ···················· | ···················· | ···················· | ···················· | ···················· | ···················· |
| R | 6385 hCoV-19/USA/MD-MDH-0341/2020\|EPI_ISL_648283\|2020-1 | ···················· | ···················· | ···················· | ···················· | ···················· | ···················· |
| R | 6386 hCoV-19/USA/MD-MDH-0344/2020\|EPI_ISL_648284\|2020-1 | ···················· | ···················· | ···················· | ···················· | ···················· | ···················· |
| R | 6387 hCoV-19/USA/MD-MDH-0345/2020\|EPI_ISL_648285\|2020-1 | ···················· | ···················· | ···················· | ···················· | ···················· | ···················· |
| R | 6388 hCoV-19/USA/MD-MDH-0347/2020\|EPI_ISL_648287\|2020-1 | ···················· | ···················· | ···················· | ···················· | ···················· | ···················· |
| R | 6389 hCoV-19/USA/MD-MDH-0348/2020\|EPI_ISL_648288\|2020-1 | ···················· | ···················· | ···················· | ···················· | ···················· | ···················· |
| R | 6390 hCoV-19/USA/MD-MDH-0350/2020\|EPI_ISL_648289\|2020-1 | ···················· | ···················· | ···················· | ···················· | ···················· | ···················· |
| R | 6391 hCoV-19/USA/MD-MDH-0351/2020\|EPI_ISL_648290\|2020-1 | ···················· | ···················· | ···················· | ···················· | ···················· | ···················· |
| R | 6392 hCoV-19/USA/MD-MDH-0352/2020\|EPI_ISL_648291\|2020-1 | ···················· | ···A················ | ···················· | ···················· | ···················· | ···················· |
| R | 6393 hCoV-19/USA/MD-MDH-0353/2020\|EPI_ISL_648292\|2020-1 | ···················· | ···················· | ···················· | ···················· | ···················· | ···················· |
| R | 6394 hCoV-19/USA/MD-MDH-0354/2020\|EPI_ISL_648293\|2020-1 | ···················· | ···················· | ···················· | ···················· | ···················· | ···················· |
| R | 6395 hCoV-19/USA/MD-MDH-0355/2020\|EPI_ISL_648294\|2020-1 | ···················· | ···A················ | ···················· | ···················· | ···················· | ···················· |
| R | 6396 hCoV-19/USA/MD-MDH-0358/2020\|EPI_ISL_648295\|2020-1 | ···················· | ···················· | ···················· | ···················· | ···················· | ···················· |
| R | 6397 hCoV-19/USA/MD-MDH-0360/2020\|EPI_ISL_648296\|2020-1 | ···················· | ···················· | ···················· | ···················· | ···················· | ···················· |
| R | 6398 hCoV-19/USA/MD-MDH-0361/2020\|EPI_ISL_648297\|2020-1 | ···················· | ·A·················· | ···················· | ···················· | ···················· | ···················· |
| R | 6399 hCoV-19/USA/MD-MDH-0363/2020\|EPI_ISL_648298\|2020-1 | ···················· | ···················· | ···················· | ···················· | ···················· | ···················· |
| R | 6400 hCoV-19/USA/MD-MDH-0364/2020\|EPI_ISL_648299\|2020-1 | ···················· | ···················· | ···················· | ···················· | ···················· | ···················· |
| R | 6401 hCoV-19/England/QEUH-B0DCF5/2020\|EPI_ISL_642009\|20 | ···················· | ···················· | ···················· | ···················· | ···················· | ···················· |
| R | 6402 hCoV-19/England/QEUH-B0EB8E/2020\|EPI_ISL_642010\|20 | ···················· | ···················· | ···················· | ···················· | ···················· | ···················· |
| R | 6403 hCoV-19/England/QEUH-B0F358/2020\|EPI_ISL_642012\|20 | ···················· | ···················· | ···················· | ···················· | ···················· | ···················· |
| R | 6404 hCoV-19/England/QEUH-B0EA09/2020\|EPI_ISL_642013\|20 | ···················· | ···················· | ···················· | ···················· | ···················· | ···················· |
| R | 6405 hCoV-19/England/QEUH-B0ECB8/2020\|EPI_ISL_642014\|20 | ···················· | ···················· | ···················· | ···················· | ···················· | ···················· |
| R | 6406 hCoV-19/England/QEUH-B0E26B/2020\|EPI_ISL_642015\|20 | ···················· | ···················· | ···················· | ···················· | ···················· | ···················· |
| R | 6407 hCoV-19/England/QEUH-B0E131/2020\|EPI_ISL_642016\|20 | ···················· | ···················· | ···················· | ···················· | ···················· | ···················· |
| R | 6408 hCoV-19/England/QEUH-B0E4ED/2020\|EPI_ISL_642017\|20 | ···················· | ···················· | ···················· | ···················· | ···················· | ···················· |
| R | 6409 hCoV-19/England/QEUH-B0E544/2020\|EPI_ISL_642018\|20 | ···················· | ···················· | ···················· | ···················· | ···················· | ···················· |
| R | 6410 hCoV-19/England/QEUH-B0E8D2/2020\|EPI_ISL_642019\|20 | ···················· | ···················· | ···················· | ···················· | ···················· | ···················· |
| R | 6411 hCoV-19/England/QEUH-B0E2F2/2020\|EPI_ISL_642020\|20 | ···················· | ···················· | ···················· | ···················· | ···················· | ···················· |
| R | 6412 hCoV-19/England/QEUH-B0E0E9/2020\|EPI_ISL_642021\|20 | ···················· | ···················· | ···················· | ···················· | ···················· | ···················· |
| R | 6413 hCoV-19/England/QEUH-B0E9B1/2020\|EPI_ISL_642022\|20 | ···················· | ···················· | ···················· | ···················· | ···················· | ···················· |
| R | 6414 hCoV-19/England/QEUH-B0E201/2020\|EPI_ISL_642023\|20 | ···················· | ···················· | ···················· | ···················· | ···················· | ···················· |
| R | 6415 hCoV-19/England/QEUH-B0DB43/2020\|EPI_ISL_642024\|20 | ···················· | ···················· | ···················· | ···················· | ···················· | ···················· |
| R | 6416 hCoV-19/England/QEUH-B0E061/2020\|EPI_ISL_642025\|20 | ···················· | ···················· | ···················· | ···················· | ···················· | ···················· |
| R | 6417 hCoV-19/England/QEUH-B0E140/2020\|EPI_ISL_642026\|20 | ···················· | ···················· | ···················· | ···················· | ···················· | ···················· |
| R | 6418 hCoV-19/England/QEUH-B0EB42/2020\|EPI_ISL_642027\|20 | ···················· | ···················· | ···················· | ···················· | ···················· | ···················· |
| R | 6419 hCoV-19/England/QEUH-B0E66F/2020\|EPI_ISL_642028\|20 | ···················· | ···················· | ···················· | ···················· | ···················· | ···················· |
| R | 6420 hCoV-19/England/QEUH-B0EA36/2020\|EPI_ISL_642029\|20 | ···················· | ···················· | ···················· | ···················· | ···················· | ···················· |
| R | 6421 hCoV-19/England/QEUH-B0FB7E/2020\|EPI_ISL_642030\|20 | ···················· | ···················· | ···················· | ···················· | ···················· | ···················· |
| R | 6422 hCoV-19/England/QEUH-B0F428/2020\|EPI_ISL_642035\|20 | ···················· | ···················· | ···················· | ···················· | ···················· | ···················· |
| R | 6423 hCoV-19/England/QEUH-B10F7E/2020\|EPI_ISL_642040\|20 | ···················· | ···················· | ···················· | ···················· | ···················· | ···················· |
| R | 6424 hCoV-19/England/QEUH-B0EC7C/2020\|EPI_ISL_642041\|20 | ···················· | ···················· | ···················· | ···················· | ···················· | ···················· |
| R | 6425 hCoV-19/England/QEUH-B0E23E/2020\|EPI_ISL_642042\|20 | ···················· | ···················· | ···················· | ···················· | ···················· | ···················· |
| R | 6426 hCoV-19/England/QEUH-B0FCA8/2020\|EPI_ISL_642043\|20 | ···················· | ···················· | ···················· | ···················· | ···················· | ···················· |
| R | 6427 hCoV-19/England/QEUH-B0E948/2020\|EPI_ISL_642044\|20 | ···················· | ···················· | ···················· | ···················· | ···················· | ···················· |
| R | 6428 hCoV-19/England/QEUH-B0E9C0/2020\|EPI_ISL_642045\|20 | ···················· | ···················· | ···················· | ···················· | ···················· | ···················· |
| R | 6429 hCoV-19/England/QEUH-B0E3E0/2020\|EPI_ISL_642046\|20 | ···················· | ···················· | ···················· | ···················· | ···················· | ···················· |
| R | 6430 hCoV-19/England/QEUH-B0F9ED/2020\|EPI_ISL_642047\|20 | ···················· | ···················· | ···················· | ···················· | ···················· | ···················· |
| R | 6431 hCoV-19/England/QEUH-B0E070/2020\|EPI_ISL_642048\|20 | ···················· | ···················· | ···················· | ···················· | ···················· | ···················· |
| R | 6432 hCoV-19/England/QEUH-B0E2A7/2020\|EPI_ISL_642049\|20 | ···················· | ···················· | ···················· | ···················· | ···················· | ···················· |
| R | 6433 hCoV-19/England/QEUH-B0F947/2020\|EPI_ISL_642050\|20 | ···················· | ···················· | ···················· | ···················· | ···················· | ···················· |
| R | 6434 hCoV-19/England/QEUH-B0DE0E/2020\|EPI_ISL_642051\|20 | ···················· | ···················· | ···················· | ···················· | ···················· | ···················· |
| R | 6435 hCoV-19/England/QEUH-B0E59F/2020\|EPI_ISL_642052\|20 | ···················· | ···················· | ···················· | ···················· | ···················· | ···················· |
| R | 6436 hCoV-19/England/QEUH-B0E4CF/2020\|EPI_ISL_642053\|20 | ···················· | ·A·················· | ···················· | ···················· | ···················· | ···A················ |
| R | 6437 hCoV-19/England/QEUH-B0E456/2020\|EPI_ISL_642054\|20 | ···················· | ···················· | ···················· | ···················· | ···················· | ···················· |
| R | 6438 hCoV-19/England/QEUH-B0DE4A/2020\|EPI_ISL_642055\|20 | ···················· | ···················· | ···················· | ···················· | ···················· | ···················· |
| R | 6439 hCoV-19/England/QEUH-B0DC31/2020\|EPI_ISL_642057\|20 | ···················· | ···················· | ···················· | ···················· | ···················· | ···················· |
| R | 6440 hCoV-19/England/QEUH-B0DAA0/2020\|EPI_ISL_642058\|20 | ···················· | ···················· | ···················· | ···················· | ···················· | ···················· |
| R | 6441 hCoV-19/England/QEUH-B0E8F0/2020\|EPI_ISL_642059\|20 | ···················· | ···················· | ···················· | ···················· | ···················· | ···················· |

| | | | | | | | | |
|---|---|---|---|---|---|---|---|---|
| R | 6442 | hCoV-19/England/QEUH-B0DE3B/2020\|EPI_ISL_642060\|20 | ···················· | ·A·················· | ···················· | ···················· | ···················· | ··A················· |
| R | 6443 | hCoV-19/England/QEUH-B0DFCF/2020\|EPI_ISL_642061\|20 | ···················· | ···················· | ···················· | ···················· | ···················· | ···················· |
| R | 6444 | hCoV-19/England/QEUH-B0DC8C/2020\|EPI_ISL_642062\|20 | ···················· | ···················· | ···················· | ···················· | ···················· | ···················· |
| R | 6445 | hCoV-19/England/QEUH-B0E562/2020\|EPI_ISL_642063\|20 | ···················· | ···················· | ···················· | ···················· | ···················· | ···················· |
| R | 6446 | hCoV-19/England/QEUH-B0EAAF/2020\|EPI_ISL_642064\|20 | ···················· | ···················· | ···················· | ···················· | ···················· | ···················· |
| R | 6447 | hCoV-19/England/QEUH-B0DFED/2020\|EPI_ISL_642065\|20 | ···················· | ···················· | ···················· | ···················· | ···················· | ···················· |
| R | 6448 | hCoV-19/England/QEUH-B0E3D1/2020\|EPI_ISL_642066\|20 | ···················· | ···················· | ···················· | ···················· | ···················· | ···················· |
| R | 6449 | hCoV-19/England/QEUH-B0EB15/2020\|EPI_ISL_642067\|20 | ···················· | ···················· | ···················· | ···················· | ···················· | ···················· |
| R | 6450 | hCoV-19/England/QEUH-B0FBBA/2020\|EPI_ISL_642068\|20 | ···················· | ···················· | ···················· | ···················· | ···················· | ···················· |
| R | 6451 | hCoV-19/England/QEUH-B0DF83/2020\|EPI_ISL_642069\|20 | ···················· | ···················· | ···················· | ···················· | ···················· | ···················· |
| R | 6452 | hCoV-19/England/QEUH-B0DE2C/2020\|EPI_ISL_642070\|20 | ···················· | ···················· | ···················· | ···················· | ···················· | ···················· |
| R | 6453 | hCoV-19/England/QEUH-B0E09E/2020\|EPI_ISL_642071\|20 | ···················· | ···················· | ···················· | ···················· | ···················· | ···················· |
| R | 6454 | hCoV-19/England/QEUH-B0F8FF/2020\|EPI_ISL_642072\|20 | ···················· | ···················· | ···················· | ···················· | ···················· | ···················· |
| R | 6455 | hCoV-19/England/QEUH-B0E1E6/2020\|EPI_ISL_642073\|20 | ···················· | ···················· | ···················· | ···················· | ···················· | ···················· |
| R | 6456 | hCoV-19/England/QEUH-B0E623/2020\|EPI_ISL_642074\|20 | ···················· | ···················· | ···················· | ···················· | ···················· | ···················· |
| R | 6457 | hCoV-19/England/QEUH-B0DDF2/2020\|EPI_ISL_642075\|20 | ···················· | ···················· | ···················· | ···················· | ···················· | ···················· |
| R | 6458 | hCoV-19/England/QEUH-B0E41A/2020\|EPI_ISL_642076\|20 | ···················· | ···················· | ···················· | ···················· | ···················· | ···················· |
| R | 6459 | hCoV-19/England/QEUH-B0DC7D/2020\|EPI_ISL_642078\|20 | ···················· | ···················· | ···················· | ···················· | ···················· | ···················· |
| R | 6460 | hCoV-19/England/QEUH-B0DEC2/2020\|EPI_ISL_642079\|20 | ···················· | ···················· | ···················· | ···················· | ···················· | ···················· |
| R | 6461 | hCoV-19/England/QEUH-B0DCE6/2020\|EPI_ISL_642080\|20 | ···················· | ···················· | ···················· | ···················· | ···················· | ···················· |
| R | 6462 | hCoV-19/England/QEUH-B0E8B4/2020\|EPI_ISL_642081\|20 | ···················· | ···················· | ···················· | ···················· | ···················· | ···················· |
| R | 6463 | hCoV-19/England/QEUH-B0E91B/2020\|EPI_ISL_642082\|20 | ···················· | ···················· | ···················· | ···················· | ···················· | ···················· |
| R | 6464 | hCoV-19/England/QEUH-B0E24D/2020\|EPI_ISL_642083\|20 | ···················· | ···················· | ···················· | ···················· | ···················· | ···················· |
| R | 6465 | hCoV-19/England/QEUH-B0E1F5/2020\|EPI_ISL_642084\|20 | ···················· | ···················· | ···················· | ···················· | ···················· | ···················· |
| R | 6466 | hCoV-19/England/QEUH-B0F8A4/2020\|EPI_ISL_642085\|20 | ···················· | ···················· | ···················· | ···················· | ···················· | ···················· |
| R | 6467 | hCoV-19/England/QEUH-B0EA54/2020\|EPI_ISL_642086\|20 | ···················· | ···················· | ···················· | ···················· | ···················· | ···················· |
| R | 6468 | hCoV-19/England/QEUH-B0E18C/2020\|EPI_ISL_642087\|20 | ···················· | ···················· | ···················· | ···················· | ···················· | ···················· |
| R | 6469 | hCoV-19/England/QEUH-B0E6C9/2020\|EPI_ISL_642088\|20 | ···················· | ···················· | ···················· | ···················· | ···················· | ···················· |
| R | 6470 | hCoV-19/England/QEUH-B0EACD/2020\|EPI_ISL_642089\|20 | ···················· | ···················· | ···················· | ···················· | ···················· | ···················· |
| R | 6471 | hCoV-19/England/QEUH-B0DDA7/2020\|EPI_ISL_642090\|20 | ···················· | ···················· | ···················· | ···················· | ···················· | ···················· |
| R | 6472 | hCoV-19/England/QEUH-B0DCD7/2020\|EPI_ISL_642091\|20 | ···················· | ···················· | ···················· | ···················· | ···················· | ···················· |
| R | 6473 | hCoV-19/England/QEUH-B0FB32/2020\|EPI_ISL_642092\|20 | ···················· | ···················· | ···················· | ···················· | ···················· | ···················· |
| R | 6474 | hCoV-19/England/QEUH-B0E492/2020\|EPI_ISL_642093\|20 | ···················· | ···················· | ···················· | ···················· | ···················· | ···················· |
| R | 6475 | hCoV-19/England/QEUH-B0FC5D/2020\|EPI_ISL_642095\|20 | ···················· | ···················· | ···················· | ···················· | ···················· | ···················· |
| R | 6476 | hCoV-19/England/QEUH-B0F9A1/2020\|EPI_ISL_642096\|20 | ···················· | ···················· | ···················· | ···················· | ···················· | ···················· |
| R | 6477 | hCoV-19/England/QEUH-B0E887/2020\|EPI_ISL_642097\|20 | ···················· | ···················· | ···················· | ···················· | ···················· | ···················· |
| R | 6478 | hCoV-19/England/QEUH-B0EBE8/2020\|EPI_ISL_642098\|20 | ···················· | ···················· | ···················· | ···················· | ···················· | ···················· |
| R | 6479 | hCoV-19/England/QEUH-B0E0CB/2020\|EPI_ISL_642099\|20 | ···················· | ···················· | ···················· | ···················· | ···················· | ···················· |
| R | 6480 | hCoV-19/England/QEUH-A8B5C8/2020\|EPI_ISL_643300\|20 | ···················· | ···················· | ···················· | ···················· | ···················· | ···················· |
| R | 6481 | hCoV-19/England/QEUH-A8C961/2020\|EPI_ISL_643301\|20 | ···················· | ···················· | ···················· | ···················· | ···················· | ···················· |
| R | 6482 | hCoV-19/England/QEUH-A8C8CE/2020\|EPI_ISL_643303\|20 | ···················· | ···················· | ···················· | ···················· | ···················· | ···················· |
| R | 6483 | hCoV-19/England/QEUH-A8BD39/2020\|EPI_ISL_643304\|20 | ···················· | ···················· | ···················· | ···················· | ···················· | ···················· |
| R | 6484 | hCoV-19/England/QEUH-A8BF8E/2020\|EPI_ISL_643305\|20 | ···················· | ···················· | ···················· | ···················· | ···················· | ···················· |
| R | 6485 | hCoV-19/England/QEUH-A8BDA2/2020\|EPI_ISL_643306\|20 | ···················· | ···················· | ···················· | ···················· | ···················· | ···················· |
| R | 6486 | hCoV-19/England/QEUH-A8B768/2020\|EPI_ISL_643307\|20 | ···················· | ···················· | ···················· | ···················· | ···················· | ···················· |
| R | 6487 | hCoV-19/England/QEUH-A8C22A/2020\|EPI_ISL_643308\|20 | ···················· | ···················· | ···················· | ···················· | ···················· | ···················· |
| R | 6488 | hCoV-19/England/QEUH-A8B452/2020\|EPI_ISL_643309\|20 | ···················· | ···················· | ···················· | ···················· | ···················· | ···················· |
| R | 6489 | hCoV-19/England/QEUH-A8C952/2020\|EPI_ISL_643310\|20 | ···················· | ···················· | ···················· | ···················· | ···················· | ···················· |
| R | 6490 | hCoV-19/England/QEUH-A8BBE4/2020\|EPI_ISL_643311\|20 | ···················· | ···················· | ···················· | ···················· | ···················· | ···················· |
| R | 6491 | hCoV-19/England/QEUH-A8C0B7/2020\|EPI_ISL_643312\|20 | ···················· | ···················· | ···················· | ···················· | ···················· | ···················· |
| R | 6492 | hCoV-19/England/QEUH-A8BA8D/2020\|EPI_ISL_643314\|20 | ···················· | ···················· | ···················· | ···················· | ···················· | ···················· |
| R | 6493 | hCoV-19/England/CAMC-B0C467/2020\|EPI_ISL_643315\|20 | ···················· | ···················· | ···················· | ···················· | ···················· | ···················· |
| R | 6494 | hCoV-19/England/CAMC-B0B073/2020\|EPI_ISL_643316\|20 | ···················· | ···················· | ···················· | ···················· | ···················· | ···················· |
| R | 6495 | hCoV-19/England/CAMC-B0CC23/2020\|EPI_ISL_643317\|20 | ···················· | ···················· | ···················· | ···················· | ···················· | ···················· |
| R | 6496 | hCoV-19/England/CAMC-B0B3C5/2020\|EPI_ISL_643318\|20 | ···················· | ···················· | ···················· | ···················· | ···················· | ···················· |
| R | 6497 | hCoV-19/England/CAMC-B0844D/2020\|EPI_ISL_643321\|20 | ···················· | ···················· | ···················· | ···················· | ···················· | ···················· |
| R | 6498 | hCoV-19/England/CAMC-B0B0CE/2020\|EPI_ISL_643322\|20 | ···················· | ···················· | ···················· | ···················· | ···················· | ···················· |
| R | 6499 | hCoV-19/England/CAMC-B0CBAE/2020\|EPI_ISL_643323\|20 | ···················· | ···················· | ···················· | ···················· | ···················· | ···················· |
| R | 6500 | hCoV-19/England/CAMC-B07AC4/2020\|EPI_ISL_643324\|20 | ···················· | ···················· | ···················· | ···················· | ···················· | ···················· |
| R | 6501 | hCoV-19/England/CAMC-B0B213/2020\|EPI_ISL_643325\|20 | ···················· | ···················· | ···················· | ···················· | ···················· | ···················· |
| R | 6502 | hCoV-19/England/CAMC-B0AF86/2020\|EPI_ISL_643339\|20 | ···················· | ·A·················· | ···················· | ···················· | ···················· | ··A················· |
| R | 6503 | hCoV-19/England/CAMC-B0B741/2020\|EPI_ISL_643340\|20 | ···················· | ···················· | ···················· | ···················· | ···················· | ···················· |
| R | 6504 | hCoV-19/England/CAMC-B08999/2020\|EPI_ISL_643342\|20 | ···················· | ·A·················· | ···················· | ···················· | ···················· | ··A················· |
| R | 6505 | hCoV-19/England/CAMC-B0B36B/2020\|EPI_ISL_643343\|20 | ···················· | ·A·················· | ···················· | ···················· | ···················· | ··A················· |
| R | 6506 | hCoV-19/England/CAMC-B0C91D/2020\|EPI_ISL_643344\|20 | ···················· | ···················· | ···················· | ···················· | ···················· | ···················· |
| R | 6507 | hCoV-19/England/CAMC-B0AD7D/2020\|EPI_ISL_643345\|20 | ···················· | ···················· | ···················· | ···················· | ···················· | ···················· |
| R | 6508 | hCoV-19/England/CAMC-B0B7BA/2020\|EPI_ISL_643346\|20 | ···················· | ···················· | ···················· | ···················· | ···················· | ···················· |
| R | 6509 | hCoV-19/England/CAMC-B0C86B/2020\|EPI_ISL_643347\|20 | ···················· | ···················· | ···················· | ···················· | ···················· | ···················· |
| R | 6510 | hCoV-19/England/CAMC-B0C898/2020\|EPI_ISL_643348\|20 | ···················· | ·A·················· | ···················· | ···················· | ···················· | ··A················· |
| R | 6511 | hCoV-19/England/CAMC-B0AFA4/2020\|EPI_ISL_643349\|20 | ···················· | ···················· | ···················· | ···················· | ···················· | ···················· |
| R | 6512 | hCoV-19/England/CAMC-B08076/2020\|EPI_ISL_643350\|20 | ···················· | ···················· | ···················· | ···················· | ····T··············· | ···················· |
| R | 6513 | hCoV-19/England/CAMC-B0B2D7/2020\|EPI_ISL_643351\|20 | ···················· | ···················· | ···················· | ···················· | ···················· | ···················· |
| R | 6514 | hCoV-19/England/CAMC-B0AA76/2020\|EPI_ISL_643352\|20 | ···················· | ···················· | ···················· | ···················· | ···················· | ···················· |
| R | 6515 | hCoV-19/England/CAMC-B0B0DD/2020\|EPI_ISL_643353\|20 | ···················· | ···················· | ···················· | ···················· | ···················· | ···················· |
| R | 6516 | hCoV-19/England/CAMC-B0B064/2020\|EPI_ISL_643354\|20 | ···················· | ···················· | ···················· | ···················· | ···················· | ···················· |
| R | 6517 | hCoV-19/England/CAMC-B07B58/2020\|EPI_ISL_643355\|20 | ···················· | ···················· | ···················· | ···················· | ···················· | ···················· |
| R | 6518 | hCoV-19/England/CAMC-B0B486/2020\|EPI_ISL_643357\|20 | ···················· | ···················· | ···················· | ···················· | ···················· | ···················· |
| R | 6519 | hCoV-19/England/CAMC-B0AD31/2020\|EPI_ISL_643358\|20 | ···················· | ···················· | ···················· | ···················· | ···················· | ···················· |
| R | 6520 | hCoV-19/England/CAMC-B084D4/2020\|EPI_ISL_643359\|20 | ···················· | ···················· | ···················· | ···················· | ···················· | ···················· |
| R | 6521 | hCoV-19/England/CAMC-B0CD6C/2020\|EPI_ISL_643360\|20 | ···················· | ···················· | ···················· | ···················· | ···················· | ···················· |

| | | | | | | | | |
|---|---|---|---|---|---|---|---|---|
| R | 6522 hCoV-19/England/CAMC-B0C85C/2020\|EPI_ISL_643361\|20 | ···················· | ···················· | ····················· | ···················· | ···················· | ···················· | |
| R | 6523 hCoV-19/England/CAMC-B0B495/2020\|EPI_ISL_643366\|20 | ···················· | ···················· | ····················· | ···················· | ···················· | ···················· | |
| R | 6524 hCoV-19/England/CAMC-B0A8C7/2020\|EPI_ISL_643370\|20 | ···················· | ···················· | ····················· | ···················· | ···················· | ···················· | |
| R | 6525 hCoV-19/England/CAMC-B0847A/2020\|EPI_ISL_643371\|20 | ···················· | ···················· | ····················· | ···················· | ···················· | ···················· | |
| R | 6526 hCoV-19/England/CAMC-B07DBC/2020\|EPI_ISL_643372\|20 | ···················· | ···················· | ····················· | ···················· | ···················· | ···················· | |
| R | 6527 hCoV-19/England/CAMC-B0CBDB/2020\|EPI_ISL_643373\|20 | ···················· | ···················· | ····················· | ···················· | ···················· | ···················· | |
| R | 6528 hCoV-19/England/CAMC-B0ABFB/2020\|EPI_ISL_643374\|20 | ···················· | ···················· | ····················· | ···················· | ···················· | ···················· | |
| R | 6529 hCoV-19/England/CAMC-B0D080/2020\|EPI_ISL_643376\|20 | ···················· | ···················· | ····················· | ···················· | ···················· | ···················· | |
| R | 6530 hCoV-19/England/CAMC-B0843E/2020\|EPI_ISL_643377\|20 | ···················· | ···················· | ····················· | ···················· | ···················· | ···················· | |
| R | 6531 hCoV-19/England/CAMC-B0C8C5/2020\|EPI_ISL_643379\|20 | ···················· | ···················· | ····················· | ···················· | ···················· | ···················· | |
| R | 6532 hCoV-19/England/CAMC-B0B2B9/2020\|EPI_ISL_643380\|20 | ···················· | ···················· | ····················· | ···················· | ···················· | ···················· | |
| R | 6533 hCoV-19/England/CAMC-B0CD3F/2020\|EPI_ISL_643381\|20 | ···················· | ···················· | ····················· | ···················· | ···················· | ···················· | |
| R | 6534 hCoV-19/England/CAMC-B07E5F/2020\|EPI_ISL_643382\|20 | ···················· | ···················· | ····················· | ···················· | ···················· | ···················· | |
| R | 6535 hCoV-19/England/CAMC-B0B1CB/2020\|EPI_ISL_643383\|20 | ···················· | ···················· | ····················· | ···················· | ···················· | ···················· | |
| R | 6536 hCoV-19/England/CAMC-B0C43A/2020\|EPI_ISL_643385\|20 | ···················· | ···················· | ····················· | ···················· | ···················· | ···················· | |
| R | 6537 hCoV-19/England/CAMC-B08577/2020\|EPI_ISL_643386\|20 | ···················· | ·A·················· | ····················· | ···················· | ···················· | ··A················· | |
| R | 6538 hCoV-19/England/CAMC-B0C84D/2020\|EPI_ISL_643387\|20 | ···················· | ···················· | ····················· | ···················· | ···················· | ···················· | |
| R | 6539 hCoV-19/England/CAMC-B0B7C9/2020\|EPI_ISL_643388\|20 | ···················· | ···················· | ····················· | ···················· | ···················· | ···················· | |
| R | 6540 hCoV-19/England/CAMC-B07AE2/2020\|EPI_ISL_643389\|20 | ···················· | ···················· | ····················· | ···················· | ···················· | ···················· | |
| R | 6541 hCoV-19/England/CAMC-B07AD3/2020\|EPI_ISL_643391\|20 | ···················· | ···················· | ····················· | ···················· | ···················· | ···················· | |
| R | 6542 hCoV-19/England/CAMC-B085A4/2020\|EPI_ISL_643392\|20 | ···················· | ·A·················· | ····················· | ···················· | ···················· | ··A················· | |
| R | 6543 hCoV-19/England/CAMC-B0B77E/2020\|EPI_ISL_643393\|20 | ···················· | ···················· | ····················· | ···················· | ·····T·············· | ···················· | |
| R | 6544 hCoV-19/England/CAMC-B0C476/2020\|EPI_ISL_643395\|20 | ···················· | ···················· | ····················· | ···················· | ···················· | ···················· | |
| R | 6545 hCoV-19/England/CAMC-B07D9E/2020\|EPI_ISL_643396\|20 | ···················· | ···················· | ····················· | ···················· | ···················· | ···················· | |
| R | 6546 hCoV-19/England/CAMC-B089A8/2020\|EPI_ISL_643397\|20 | ···················· | ···················· | ····················· | ···················· | ···················· | ···················· | |
| R | 6547 hCoV-19/England/CAMC-B0B732/2020\|EPI_ISL_643398\|20 | ···················· | ···················· | ····················· | ···················· | ···················· | ···················· | |
| R | 6548 hCoV-19/England/CAMC-B0AE89/2020\|EPI_ISL_643399\|20 | ···················· | ···················· | ····················· | ···················· | ···················· | ···················· | |
| R | 6549 hCoV-19/USA/NM-NMDOH-2020329989/2020\|EPI_ISL_64460 | ···················· | ···················· | ····················· | ···················· | ···················· | ···················· | |
| R | 6550 hCoV-19/USA/NM-NMDOH-2020329992/2020\|EPI_ISL_64460 | ···················· | ···················· | ····················· | ···················· | ···················· | ···················· | |
| R | 6551 hCoV-19/USA/NM-NMDOH-2020330052/2020\|EPI_ISL_64460 | ···················· | ···················· | ····················· | ···················· | ···················· | ···················· | |
| R | 6552 hCoV-19/USA/NM-NMDOH-2020330065/2020\|EPI_ISL_64460 | ···················· | ···················· | ····················· | ···················· | ···················· | ···················· | |
| R | 6553 hCoV-19/USA/NM-NMDOH-2020330134/2020\|EPI_ISL_64460 | ···················· | ···················· | ····················· | ···················· | ···················· | ···················· | |
| R | 6554 hCoV-19/USA/NM-NMDOH-2020330165/2020\|EPI_ISL_64460 | ···················· | ···················· | ····················· | ···················· | ···················· | ···················· | |
| R | 6555 hCoV-19/USA/NM-NMDOH-2020330167/2020\|EPI_ISL_64460 | ···················· | ···················· | ····················· | ···················· | ···················· | ···················· | |
| R | 6556 hCoV-19/USA/NM-NMDOH-2020330317/2020\|EPI_ISL_64460 | ···················· | ·············A······ | ····················· | ···················· | ···················· | ···················· | |
| R | 6557 hCoV-19/USA/NM-NMDOH-2020330343/2020\|EPI_ISL_64460 | ···················· | ···················· | ····················· | ···················· | ···················· | ···················· | |
| R | 6558 hCoV-19/USA/NM-NMDOH-2020330367/2020\|EPI_ISL_64460 | ···················· | ···················· | ····················· | ···················· | ···················· | ···················· | |
| R | 6559 hCoV-19/Belgium/ULG-10765/2020\|EPI_ISL_644611\|2020 | ···················· | ···················· | ····················· | ···················· | ···················· | ···················· | |
| R | 6561 hCoV-19/Belgium/ULG-10767/2020\|EPI_ISL_644613\|2020 | ···················· | ···················· | ····················· | ···················· | ···················· | ···················· | |
| R | 6563 hCoV-19/Belgium/ULG-10769/2020\|EPI_ISL_644615\|2020 | ···················· | ···················· | ····················· | ···················· | ···················· | ···················· | |
| R | 6564 hCoV-19/Belgium/ULG-10770/2020\|EPI_ISL_644616\|2020 | ···················· | ···A················ | ····················· | ···················· | ···················· | ···················· | |
| R | 6566 hCoV-19/Belgium/ULG-10773/2020\|EPI_ISL_644619\|2020 | ···················· | ···················· | ····················· | ···················· | ···················· | ···················· | |
| R | 6569 hCoV-19/Belgium/ULG-10776/2020\|EPI_ISL_644622\|2020 | ···················· | ···················· | ····················· | ···················· | ···················· | ···················· | |
| R | 6570 hCoV-19/Belgium/ULG-10778/2020\|EPI_ISL_644624\|2020 | ···················· | ···················· | ····················· | ···················· | ···················· | ···················· | |
| R | 6571 hCoV-19/Belgium/ULG-10779/2020\|EPI_ISL_644625\|2020 | ···················· | ···················· | ····················· | ···················· | ···················· | ···················· | |
| R | 6573 hCoV-19/Malaysia/IMR_CV140329/2020\|EPI_ISL_644672\| | ···················· | ·A·················· | ····················· | ···················· | ···················· | ···················· | |
| R | 6574 hCoV-19/France/NAQ-SC280/2020\|EPI_ISL_644673\|2020- | ···················· | ···················· | ····················· | ···················· | ···················· | ···················· | |
| R | 6575 hCoV-19/France/NAQ-SC281/2020\|EPI_ISL_644674\|2020- | ···················· | ···················· | ····················· | ···················· | ···················· | ···················· | |
| R | 6576 hCoV-19/France/NAQ-SC282/2020\|EPI_ISL_644675\|2020- | ···················· | ···················· | ····················· | ···················· | ···················· | ···················· | |
| R | 6577 hCoV-19/France/NAQ-SC283/2020\|EPI_ISL_644676\|2020- | ···················· | ···················· | ····················· | ···················· | ···················· | ···················· | |
| R | 6578 hCoV-19/France/NAQ-SC284/2020\|EPI_ISL_644677\|2020- | ···················· | ···················· | ····················· | ···················· | ···················· | ···················· | |
| R | 6579 hCoV-19/France/NAQ-SC288/2020\|EPI_ISL_644678\|2020- | ···················· | ···················· | ····················· | ···················· | ···················· | ···················· | |
| R | 6580 hCoV-19/France/NAQ-SC289/2020\|EPI_ISL_644679\|2020- | ···················· | ···················· | ····················· | ···················· | ···················· | ···················· | |
| R | 6581 hCoV-19/France/NAQ-SC302/2020\|EPI_ISL_644680\|2020- | ···················· | ···················· | ····················· | ···················· | ···················· | ···················· | |
| R | 6582 hCoV-19/France/OCC-SC463/2020\|EPI_ISL_644681\|2020- | ···················· | ···················· | ····················· | ···················· | ···················· | ···················· | |
| R | 6583 hCoV-19/France/OCC-SC466/2020\|EPI_ISL_644682\|2020- | ···················· | ···················· | ····················· | ···················· | ···················· | ···················· | |
| R | 6585 hCoV-19/France/OCC-SC468/2020\|EPI_ISL_644684\|2020- | ···················· | ···················· | ····················· | ···················· | ···················· | ···················· | |
| R | 6586 hCoV-19/France/OCC-SC471/2020\|EPI_ISL_644685\|2020- | ···················· | ···················A | ····················· | ···················· | ···················· | ···················· | 1 |
| R | 6587 hCoV-19/France/OCC-SC472/2020\|EPI_ISL_644686\|2020- | ···················· | ···················A | ····················· | ···················· | ···················· | ···················· | 1 |
| R | 6588 hCoV-19/France/OCC-SC474/2020\|EPI_ISL_644687\|2020- | ···················· | ·········A·········· | ····················· | ···················· | ···················· | ···················· | |
| R | 6589 hCoV-19/France/OCC-SC483/2020\|EPI_ISL_644688\|2020- | ···················· | ···················· | ····················· | ···················· | ···················· | ···················· | |
| R | 6590 hCoV-19/France/OCC-SC486/2020\|EPI_ISL_644689\|2020- | ···················· | ···················· | ····················· | ···················· | ···················· | ···················· | |
| R | 6591 hCoV-19/France/OCC-SC487/2020\|EPI_ISL_644690\|2020- | ···················· | ···················· | ····················· | ···················· | ···················· | ···················· | |
| R | 6592 hCoV-19/France/OCC-SC489/2020\|EPI_ISL_644691\|2020- | ···················· | ···················· | ····················· | ···················· | ···················· | ···················· | |
| R | 6593 hCoV-19/France/OCC-SC493/2020\|EPI_ISL_644692\|2020- | ···················· | ···················· | ····················· | ···················· | ···················· | ···················· | |
| R | 6594 hCoV-19/France/OCC-SC527/2020\|EPI_ISL_644697\|2020- | ···················· | ···················· | ····················· | ···················· | ···················· | ···················· | |
| R | 6595 hCoV-19/France/OCC-SC529/2020\|EPI_ISL_644698\|2020- | ···················· | ···················A | ····················· | ···················· | ···················· | ···················· | 1 |
| R | 6596 hCoV-19/England/ALDP-B1C033/2020\|EPI_ISL_645922\|20 | ···················· | ···················· | ····················· | ···················· | ···················· | ···················· | |
| R | 6597 hCoV-19/England/CAMC-B22076/2020\|EPI_ISL_645923\|20 | ···················· | ···················· | ····················· | ···················· | ···················· | ···················· | |
| R | 6598 hCoV-19/England/ALDP-B1DB9B/2020\|EPI_ISL_645924\|20 | ···················· | ·A·················· | ····················· | ···················· | ···················· | ···················· | |
| R | 6599 hCoV-19/England/CAMC-B2239B/2020\|EPI_ISL_645925\|20 | ···················· | ···················· | ····················· | ···················· | ···················· | ···················· | |
| R | 6600 hCoV-19/England/CAMC-B2236E/2020\|EPI_ISL_645926\|20 | ···················· | ···················· | ····················· | ···················· | ···················· | ···················· | |
| R | 6601 hCoV-19/England/CAMC-B220A3/2020\|EPI_ISL_645927\|20 | ···················· | ···················· | ····················· | ···················· | ···················· | ···················· | |
| R | 6602 hCoV-19/England/ALDP-B1F8DE/2020\|EPI_ISL_645928\|20 | ···················· | ···················· | ····················· | ···················· | ···················· | ···················· | |
| R | 6603 hCoV-19/England/ALDP-B1B878/2020\|EPI_ISL_645929\|20 | ···················· | ···················· | ····················· | ···················· | ···················· | ···················· | |
| R | 6604 hCoV-19/England/CAMC-B222CB/2020\|EPI_ISL_645930\|20 | ···················· | ···················· | ····················· | ···················· | ···················· | ···················· | |
| R | 6605 hCoV-19/England/ALDP-B1C042/2020\|EPI_ISL_645931\|20 | ···················· | ···················· | ····················· | ···················· | ···················· | ···················· | |
| R | 6606 hCoV-19/England/ALDP-B1ADB6/2020\|EPI_ISL_645932\|20 | ···················· | ···················· | ····················· | ···················· | ···················· | ···················· | |
| R | 6607 hCoV-19/England/ALDP-B1C482/2020\|EPI_ISL_645933\|20 | ···················· | ···················· | ····················· | ···················· | ···················· | ···················· | |
| R | 6608 hCoV-19/England/ALDP-B1A3A5/2020\|EPI_ISL_645934\|20 | ···················· | ···················· | ····················· | ···················· | ···················· | ··A················· | |

| | | | | | | | | |
|---|---|---|---|---|---|---|---|---|
| R | 6609 | hCoV-19/England/ALDP-B1B1D7/2020\|EPI_ISL_645935\|20 | ···················· | ···················· | ···················· | ···················· | ···················· | ···················· |
| R | 6610 | hCoV-19/England/ALDP-B1C491/2020\|EPI_ISL_645936\|20 | ···················· | ···················· | ···················· | ···················· | ···················· | ···················· |
| R | 6611 | hCoV-19/England/ALDP-B1AF0B/2020\|EPI_ISL_645937\|20 | ···················· | ···················· | ···················· | ···················· | ···················· | ···················· |
| R | 6612 | hCoV-19/England/ALDP-B1D560/2020\|EPI_ISL_645938\|20 | ···················· | ···················· | ···················· | ···················· | ···················· | ···················· |
| R | 6613 | hCoV-19/England/ALDP-B1C16D/2020\|EPI_ISL_645939\|20 | ···················· | ···················· | ···················· | ···················· | ···················· | ···················· |
| R | 6614 | hCoV-19/England/ALDP-B1ACB9/2020\|EPI_ISL_645940\|20 | ···················· | ···················· | ···················· | ···················· | ···················· | ···················· |
| R | 6615 | hCoV-19/England/ALDP-B1F81A/2020\|EPI_ISL_645941\|20 | ···················· | ···················· | ···················· | ···················· | ···················· | ···················· |
| R | 6616 | hCoV-19/England/ALDP-B1ACE6/2020\|EPI_ISL_645942\|20 | ···················· | ···················· | ···················· | ···················· | ···················· | ···················· |
| R | 6617 | hCoV-19/England/ALDP-B1F57D/2020\|EPI_ISL_645943\|20 | ···················· | ···················· | ···················· | ···················· | ···················· | ···················· |
| R | 6618 | hCoV-19/England/ALDP-B1F5F5/2020\|EPI_ISL_645944\|20 | ···················· | ···················· | ···················· | ···················· | ···················· | ···················· |
| R | 6619 | hCoV-19/England/CAMC-B222AD/2020\|EPI_ISL_645945\|20 | ···················· | ···················· | ···················· | ···················· | ···················· | ···················· |
| R | 6620 | hCoV-19/England/ALDP-B1F777/2020\|EPI_ISL_645946\|20 | ···················· | ···················· | ···················· | ···················· | ···················· | ···················· |
| R | 6621 | hCoV-19/England/ALDP-B1F829/2020\|EPI_ISL_645947\|20 | ···················· | ···················· | ···················· | ···················· | ···················· | ···················· |
| R | 6622 | hCoV-19/England/ALDP-B1C0AC/2020\|EPI_ISL_645948\|20 | ···················· | ···················· | ···················· | ···················· | ···················· | ···················· |
| R | 6623 | hCoV-19/England/CAMC-B21D07/2020\|EPI_ISL_645949\|20 | ···················· | ···················· | ···················· | ···················· | ···················· | ···················· |
| R | 6624 | hCoV-19/England/ALDP-B1B210/2020\|EPI_ISL_645950\|20 | ···················· | ···················· | ···················· | ···················· | ···················· | ···················· |
| R | 6625 | hCoV-19/England/ALDP-B1AEC2/2020\|EPI_ISL_645951\|20 | ···················· | ···················· | ···················· | ···················· | ···················· | ···················· |
| R | 6626 | hCoV-19/England/ALDP-B1AE3B/2020\|EPI_ISL_645952\|20 | ···················· | ···················· | ···················· | ···················· | ···················· | ···················· |
| R | 6627 | hCoV-19/England/ALDP-B1F5AA/2020\|EPI_ISL_645954\|20 | ···················· | ···················· | ···················· | ···················· | ···················· | ···················· |
| R | 6628 | hCoV-19/England/ALDP-B1DBB9/2020\|EPI_ISL_645955\|20 | ···················· | ···················· | ···················· | ···················· | ···················· | ···················· |
| R | 6629 | hCoV-19/England/ALDP-B1F56E/2020\|EPI_ISL_645957\|20 | ····G··············· | ···················· | ···················· | ···················· | ···················· | ···················· |
| R | 6630 | hCoV-19/England/CAMC-B223D7/2020\|EPI_ISL_645958\|20 | ···················· | ···················· | ···················· | ···················· | ···················· | ···················· |
| R | 6631 | hCoV-19/England/ALDP-B1BD4C/2020\|EPI_ISL_645960\|20 | ···················· | ···················· | ···················· | ···················· | ···················· | ···················· |
| R | 6632 | hCoV-19/England/ALDP-B1ADD4/2020\|EPI_ISL_645961\|20 | ···················· | ···················· | ···················· | ···················· | ···················· | ···················· |
| R | 6633 | hCoV-19/England/CAMC-B21FD4/2020\|EPI_ISL_645962\|20 | ···················· | ···················· | ···················· | ···················· | ···················· | ···················· |
| R | 6634 | hCoV-19/England/ALDP-B1B24D/2020\|EPI_ISL_645963\|20 | ···················· | ···················· | ···················· | ···················· | ···················· | ···················· |
| R | 6635 | hCoV-19/England/ALDP-B1D5CA/2020\|EPI_ISL_645964\|20 | ···················· | ···················· | ···················· | ···················· | ···················· | ···················· |
| R | 6636 | hCoV-19/England/ALDP-B1D515/2020\|EPI_ISL_645965\|20 | ···················· | ···················· | ···················· | ···················· | ···················· | ···················· |
| R | 6637 | hCoV-19/England/ALDP-B1BD3D/2020\|EPI_ISL_645966\|20 | ···················· | ···················· | ···················· | ···················· | ···················· | ···················· |
| R | 6638 | hCoV-19/England/ALDP-B1A2B7/2020\|EPI_ISL_645967\|20 | ···················· | ···················· | ···················· | ···················· | ···················· | ···················· |
| R | 6639 | hCoV-19/England/CAMC-B22313/2020\|EPI_ISL_645968\|20 | ···················· | ···················· | ···················· | ···················· | ···················· | ···················· |
| R | 6640 | hCoV-19/England/ALDP-B1AE86/2020\|EPI_ISL_645969\|20 | ···················· | ···················· | ···················· | ···················· | ···················· | ···················· |
| R | 6641 | hCoV-19/England/CAMC-B22049/2020\|EPI_ISL_645970\|20 | ···················· | ···················· | ···················· | ···················· | ···················· | ···················· |
| R | 6642 | hCoV-19/England/ALDP-B1ADF2/2020\|EPI_ISL_645971\|20 | ···················· | ···················· | ···················· | ···················· | ···················· | ···················· |
| R | 6643 | hCoV-19/England/ALDP-B1D5F7/2020\|EPI_ISL_645972\|20 | ···················· | ···················· | ···················· | ···················· | ···················· | ···················· |
| R | 6644 | hCoV-19/England/ALDP-B1AE2C/2020\|EPI_ISL_645973\|20 | ···················· | ···················· | ···················· | ···················· | ···················· | ···················· |
| R | 6645 | hCoV-19/England/ALDP-B1B2A7/2020\|EPI_ISL_645974\|20 | ···················· | ···················· | ···················· | ···················· | ···················· | ···················· |
| R | 6646 | hCoV-19/England/ALDP-B1AD6B/2020\|EPI_ISL_645975\|20 | ···················· | ···················· | ···················· | ···················· | ···················· | ···················· |
| R | 6647 | hCoV-19/England/ALDP-B1DB13/2020\|EPI_ISL_645976\|20 | ···················· | ···················· | ···················· | ···················· | ···················· | ···················· |
| R | 6648 | hCoV-19/England/ALDP-B1A33C/2020\|EPI_ISL_645977\|20 | ···················· | ···················· | ···················· | ···················· | ···················· | ···················· |
| R | 6649 | hCoV-19/England/CAMC-B2202B/2020\|EPI_ISL_645980\|20 | ···················· | ···················· | ···················· | ···················· | ···················· | ···················· |
| R | 6650 | hCoV-19/England/CAMC-B22094/2020\|EPI_ISL_645981\|20 | ···················· | ···················· | ···················· | ···················· | ···················· | ···················· |
| R | 6651 | hCoV-19/England/ALDP-B1B7F3/2020\|EPI_ISL_645982\|20 | ···················· | ···················· | ···················· | ···················· | ···················· | ···················· |
| R | 6652 | hCoV-19/England/CAMC-B21BEF/2020\|EPI_ISL_645983\|20 | ···················· | ···················· | ···················· | ···················· | ···················· | ···················· |
| R | 6653 | hCoV-19/England/ALDP-B1A32D/2020\|EPI_ISL_645984\|20 | ···················· | ···················· | ···················· | ···················· | ···················· | ···················· |
| R | 6654 | hCoV-19/England/ALDP-B1F8FC/2020\|EPI_ISL_645985\|20 | ···················· | ···················· | ···················· | ···················· | ···················· | ···················· |
| R | 6655 | hCoV-19/England/ALDP-B1AC7D/2020\|EPI_ISL_645986\|20 | ···················· | ···················· | ···················· | ···················· | ···················· | ···················· |
| R | 6656 | hCoV-19/England/CAMC-B2261A/2020\|EPI_ISL_645987\|20 | ···················· | ·A·················· | ···················· | ···················· | ···················· | ··A················· |
| R | 6657 | hCoV-19/England/QEUH-B0DFA1/2020\|EPI_ISL_642031\|20 | ···················· | ···················· | ···················· | ···················· | ···················· | ···················· |
| R | 6658 | hCoV-19/England/QEUH-B0E939/2020\|EPI_ISL_642032\|20 | ···················· | ···················· | ···················· | ···················· | ···················· | ···················· |
| R | 6659 | hCoV-19/England/QEUH-B0E007/2020\|EPI_ISL_642034\|20 | ···················· | ···················· | ···················· | ···················· | ···················· | ···················· |
| R | 6660 | hCoV-19/England/QEUH-B0EC8B/2020\|EPI_ISL_642036\|20 | ···················· | ···················· | ···················· | ···················· | ···················· | ···················· |
| R | 6661 | hCoV-19/England/QEUH-B0E25C/2020\|EPI_ISL_642037\|20 | ···················· | ···················· | ···················· | ···················· | ···················· | ···················· |
| R | 6662 | hCoV-19/England/QEUH-B0E5EA/2020\|EPI_ISL_642038\|20 | ···················· | ···················· | ···················· | ···················· | ······T············· | ···················· |
| R | 6663 | hCoV-19/England/QEUH-A8C6A6/2020\|EPI_ISL_643200\|20 | ···················· | ···················· | ···················· | ···················· | ···················· | ···················· |
| R | 6664 | hCoV-19/England/QEUH-A8C794/2020\|EPI_ISL_643201\|20 | ···················· | ···················· | ···················· | ···················· | ···················· | ···················· |
| R | 6665 | hCoV-19/England/QEUH-A8BD66/2020\|EPI_ISL_643202\|20 | ···················· | ···················· | ···················· | ···················· | ···················· | ···················· |
| R | 6666 | hCoV-19/England/QEUH-A8B9CC/2020\|EPI_ISL_643203\|20 | ···················· | ···················· | ···················· | ···················· | ···················· | ···················· |
| R | 6667 | hCoV-19/England/QEUH-A8BE36/2020\|EPI_ISL_643204\|20 | ···················· | ···················· | ···················· | ···················· | ···················· | ···················· |
| R | 6668 | hCoV-19/England/QEUH-A8BF7F/2020\|EPI_ISL_643205\|20 | ···················· | ···················· | ···················· | ···················· | ···················· | ···················· |
| R | 6669 | hCoV-19/England/QEUH-A8CA6E/2020\|EPI_ISL_643206\|20 | ···················· | ···················· | ···················· | ···················· | ···················· | ···················· |
| R | 6670 | hCoV-19/England/QEUH-A8BF24/2020\|EPI_ISL_643207\|20 | ···················· | ···················· | ···················· | ···················· | ···················· | ···················· |
| R | 6671 | hCoV-19/England/QEUH-A8BD1B/2020\|EPI_ISL_643208\|20 | ···················· | ···················· | ···················· | ···················· | ···················· | ···················· |
| R | 6672 | hCoV-19/England/QEUH-A8BAAB/2020\|EPI_ISL_643209\|20 | ···················· | ···················· | ···················· | ···················· | ···················· | ···················· |
| R | 6673 | hCoV-19/England/QEUH-A8C5F4/2020\|EPI_ISL_643210\|20 | ···················· | ···················· | ···················· | ···················· | ···················· | ···················· |
| R | 6674 | hCoV-19/England/QEUH-A8C1A5/2020\|EPI_ISL_643211\|20 | ···················· | ···················· | ···················· | ···················· | ···················· | ···················· |
| R | 6675 | hCoV-19/England/QEUH-A8CA40/2020\|EPI_ISL_643212\|20 | ···················· | ···················· | ···················· | ···················· | ···················· | ···················· |
| R | 6676 | hCoV-19/England/QEUH-A8B610/2020\|EPI_ISL_643214\|20 | ···················· | ·A·················· | ···················· | ···················· | ···················· | ···················· |
| R | 6677 | hCoV-19/England/QEUH-A8BAF6/2020\|EPI_ISL_643215\|20 | ···················· | ···················· | ···················· | ···················· | ···················· | ···················· |
| R | 6678 | hCoV-19/England/QEUH-A8B4CB/2020\|EPI_ISL_643216\|20 | ···················· | ···················· | ···················· | ···················· | ···················· | ···················· |
| R | 6679 | hCoV-19/England/QEUH-A8B786/2020\|EPI_ISL_643217\|20 | ···················· | ···················· | ···················· | ···················· | ···················· | ···················· |
| R | 6680 | hCoV-19/England/QEUH-A8BB7B/2020\|EPI_ISL_643218\|20 | ···················· | ···················· | ···················· | ···················· | ···················· | ···················· |
| R | 6681 | hCoV-19/England/QEUH-A8C1B4/2020\|EPI_ISL_643219\|20 | ···················· | ···················· | ···················· | ···················· | ···················· | ···················· |
| R | 6682 | hCoV-19/England/QEUH-A8B6D4/2020\|EPI_ISL_643220\|20 | ···················· | ···················· | ···················· | ···················· | ···················· | ···················· |
| R | 6683 | hCoV-19/England/QEUH-A8C10F/2020\|EPI_ISL_643221\|20 | ···················· | ···················· | ···················· | ···················· | ···················· | ···················· |
| R | 6684 | hCoV-19/England/QEUH-A8BDDF/2020\|EPI_ISL_643222\|20 | ···················· | ···················· | ···················· | ···················· | ···················· | ···················· |
| R | 6685 | hCoV-19/England/QEUH-A8C4E8/2020\|EPI_ISL_643223\|20 | ···················· | ···················· | ···················· | ···················· | ···················· | ···················· |
| R | 6686 | hCoV-19/England/QEUH-A8C6D3/2020\|EPI_ISL_643224\|20 | ···················· | ···················· | ···················· | ···················· | ···················· | ···················· |
| R | 6687 | hCoV-19/England/QEUH-A8C6F1/2020\|EPI_ISL_643225\|20 | ···················· | ···················· | ···················· | ···················· | ···················· | ···················· |
| R | 6688 | hCoV-19/England/QEUH-A8B874/2020\|EPI_ISL_643226\|20 | ···················· | ···················· | ···················· | ···················· | ···················· | ···················· |

R 6689 hCoV-19/England/CAMC-B0ABEC/2020|EPI_ISL_643362|20 ···················· ···················· ···················· ···················· ···················· ····················
R 6690 hCoV-19/England/CAMC-B0AB19/2020|EPI_ISL_643363|20 ···················· ···················· ···················· ···················· ···················· ····················
R 6691 hCoV-19/England/CAMC-B08058/2020|EPI_ISL_643364|20 ···················· ···················· ···················· ···················· ···················· ····················
R 6692 hCoV-19/England/CAMC-B07AF1/2020|EPI_ISL_643365|20 ···················· ···················· ···················· ···················· ···················· ····················
R 6693 hCoV-19/England/CAMC-B0CC9C/2020|EPI_ISL_643367|20 ···················· ···················· ···················· ···················· ···················· ····················
R 6694 hCoV-19/England/CAMC-B084A7/2020|EPI_ISL_643368|20 ···················· ···················· ···················· ···················· ···················· ····················
R 6695 hCoV-19/England/CAMC-B0851D/2020|EPI_ISL_643369|20 ···················· ···················· ···················· ···················· ···················· ····················
R 6696 hCoV-19/France/PAC-MEPHI-2112/2020|EPI_ISL_644500| ···················· ···················· ···················· ···················· ···················· ····················
R 6697 hCoV-19/France/PAC-MEPHI-2117/2020|EPI_ISL_644502| ···················· ···················· ···················· ···················· ···················· ····················
R 6698 hCoV-19/France/PAC-MEPHI-2153/2020|EPI_ISL_644514| ···················· ···················· ···················· ···················· ···················· ····················
R 6699 hCoV-19/France/PAC-MEPHI-2154/2020|EPI_ISL_644515| ···················· ···················· ···················· ···················· ···················· ····················
R 6700 hCoV-19/France/PAC-MEPHI-2155/2020|EPI_ISL_644516| ···················· ···················· ···················· ···················· ···················· ····················
R 6701 hCoV-19/France/PAC-MEPHI-2157/2020|EPI_ISL_644517| ···················· ···················· ···················· ···················· ···················· ····················
R 6702 hCoV-19/France/PAC-MEPHI-2161/2020|EPI_ISL_644518| ···················· ···················· ···················· ···················· ···················· ····················
R 6703 hCoV-19/France/PAC-MEPHI-2165/2020|EPI_ISL_644519| ···T················ ···················· ···················· ···················· ···················· ····················
R 6704 hCoV-19/France/PAC-MEPHI-2166/2020|EPI_ISL_644520| ···················· ···················· ···················· ···················· ···················· ····················
R 6705 hCoV-19/France/PAC-MEPHI-2172/2020|EPI_ISL_644521| ···················· ···················· ···················· ···················· ···················· ····················
R 6706 hCoV-19/France/PAC-MEPHI-2173/2020|EPI_ISL_644522| ···················· ···················· ···················· ···················· ···················· ····················
R 6707 hCoV-19/France/PAC-MEPHI-2180/2020|EPI_ISL_644523| ···················· ···················· ···················· ···················· ···················· ····················
R 6708 hCoV-19/France/PAC-MEPHI-2181/2020|EPI_ISL_644524| ···················· ···················· ···················· ···················· ···················· ····················
R 6709 hCoV-19/France/PAC-MEPHI-2195/2020|EPI_ISL_644525| ···················· ···················· ···················· ···················· ···················· ····················
R 6710 hCoV-19/France/PAC-MEPHI-2207/2020|EPI_ISL_644526| ···················· ···················· ···················· ···················· ···················· ····················
R 6711 hCoV-19/France/PAC-MEPHI-2211/2020|EPI_ISL_644527| ···················· ···················· ···················· ···················· ···················· ····················
R 6712 hCoV-19/France/PAC-MEPHI-2221/2020|EPI_ISL_644528| ···················· ···················· ···················· ···················· ···················· ····················
R 6713 hCoV-19/France/PAC-MEPHI-2227/2020|EPI_ISL_644529| ···················· ···················· ···················· ···················· ···················· ····················
R 6714 hCoV-19/France/PAC-MEPHI-2233/2020|EPI_ISL_644530| ···················· ···················· ···················· ···················· ···················· ····················
R 6715 hCoV-19/France/PAC-MEPHI-2272/2020|EPI_ISL_644531| ···················· ···················· ···················· ···················· ···················· ····················
R 6716 hCoV-19/France/PAC-MEPHI-2281/2020|EPI_ISL_644532| ···················· ····A··············· ···················· ···················· ···················· ····················
R 6717 hCoV-19/France/PAC-MEPHI-2288/2020|EPI_ISL_644533| ···················· ···················· ···················· ···················· ···················· ····················
R 6718 hCoV-19/France/PAC-MEPHI-2289/2020|EPI_ISL_644534| ···················· ···················· ···················· ···················· ···················· ····················
R 6719 hCoV-19/France/PAC-MEPHI-2295/2020|EPI_ISL_644535| ···················· ···················· ···················· ···················· ···················· ····················
R 6720 hCoV-19/France/PAC-MEPHI-2304/2020|EPI_ISL_644536| ···················· ···················· ···················· ···················· ···················· ····················
R 6721 hCoV-19/France/PAC-MEPHI-2308/2020|EPI_ISL_644537| ···················· ···················· ···················· ···················· ···················· ····················
R 6722 hCoV-19/France/PAC-MEPHI-2320/2020|EPI_ISL_644538| ···················· ···················· ···················· ···················· ···················· ····················
R 6723 hCoV-19/France/PAC-MEPHI-2321/2020|EPI_ISL_644539| ···················· ···················· ···················· ···················· ···················· ····················
R 6724 hCoV-19/France/PAC-MEPHI-2326/2020|EPI_ISL_644540| ···················· ···················· ···················· ···················· ···················· ····················
R 6725 hCoV-19/France/PAC-MEPHI-2341/2020|EPI_ISL_644541| ···················· ···················· ···················· ···················· ···················· ····················
R 6726 hCoV-19/France/PAC-MEPHI-2344/2020|EPI_ISL_644542| ···················· ···················· ···················· ···················· ···················· ····················
R 6727 hCoV-19/France/PAC-MEPHI-2368/2020|EPI_ISL_644543| ···················· ···················· ···················· ···················· ···················· ····················
R 6728 hCoV-19/France/PAC-MEPHI-2369/2020|EPI_ISL_644544| ···················· ···················· ···················· ···················· ···················· ····················
R 6729 hCoV-19/France/PAC-MEPHI-2371/2020|EPI_ISL_644545| ···················· ···················· ···················· ···················· ···················· ····················
R 6730 hCoV-19/France/PAC-MEPHI-2374/2020|EPI_ISL_644546| ···················· ···················· ···················· ···················· ···················· ····················
R 6731 hCoV-19/France/PAC-MEPHI-2375/2020|EPI_ISL_644547| ···················· ···················· ···················· ···················· ···················· ····················
R 6732 hCoV-19/France/PAC-MEPHI-2376/2020|EPI_ISL_644548| ···················· ···················· ···················· ···················· ···················· ····················
R 6733 hCoV-19/France/PAC-MEPHI-2382/2020|EPI_ISL_644549| ···················· ···················· ···················· ···················· ···················· ····················
R 6734 hCoV-19/France/PAC-MEPHI-2385/2020|EPI_ISL_644550| ···················· ···················· ···················· ···················· ···················· ····················
R 6735 hCoV-19/France/PAC-MEPHI-2388/2020|EPI_ISL_644551| ···················· ···················· ···················· ···················· ···················· ····················
R 6736 hCoV-19/France/PAC-MEPHI-2389/2020|EPI_ISL_644552| ···················· ···················· ···················· ···················· ···················· ····················
R 6737 hCoV-19/France/PAC-MEPHI-2390/2020|EPI_ISL_644553| ···················· ···················· ···················· ···················· ···················· ····················
R 6738 hCoV-19/France/PAC-MEPHI-2393/2020|EPI_ISL_644554| ···················· ···················· ···················· ···················· ···················· ····················
R 6739 hCoV-19/France/PAC-MEPHI-2394/2020|EPI_ISL_644555| ···················· ···················· ···················· ···················· ···················· ····················
R 6740 hCoV-19/France/PAC-MEPHI-2400/2020|EPI_ISL_644556| ···················· ···················· ···················· ···················· ···················· ····················
R 6741 hCoV-19/France/PAC-MEPHI-2401/2020|EPI_ISL_644557| ···················· ···················· ···················· ···················· ···················· ····················
R 6742 hCoV-19/France/OCC-SC496/2020|EPI_ISL_644693|2020- ···················· ···················· ···················· ···················· ···················· ····················
R 6743 hCoV-19/France/OCC-SC503/2020|EPI_ISL_644694|2020- ···················· ···················· ···················· ···················· ···················· ····················
R 6744 hCoV-19/France/OCC-SC504/2020|EPI_ISL_644695|2020- ···················· ···················· ···················· ···················· ···················· ····················
R 6745 hCoV-19/France/OCC-SC516/2020|EPI_ISL_644696|2020- ···················· ···················· ···················· ···················· ···················· ····················
R 6746 hCoV-19/France/OCC-SC530/2020|EPI_ISL_644699|2020- ···················· ···················· ···················· ···················· ···················· ····················
R 6747 hCoV-19/England/ALDP-B1B5AE/2020|EPI_ISL_645823|20 ···················· ···················· ···················· ···················· ···················· ····················
R 6748 hCoV-19/England/ALDP-B19F48/2020|EPI_ISL_645824|20 ···················· ···················· ···················· ···················· ···················· ····················
R 6749 hCoV-19/England/ALDP-B1B23E/2020|EPI_ISL_645825|20 ···················· ···················· ···················· ···················· ···················· ····················
R 6750 hCoV-19/England/CAMC-B223B9/2020|EPI_ISL_645826|20 ···················· ···················· ···················· ···················· ···················· ····················
R 6751 hCoV-19/England/ALDP-B1B8B4/2020|EPI_ISL_645827|20 ···················· ···················· ···················· ···················· ···················· ····················
R 6752 hCoV-19/England/CAMC-B22058/2020|EPI_ISL_645828|20 ···················· ···················· ···················· ···················· ···················· ····················
R 6753 hCoV-19/England/CAMC-B22146/2020|EPI_ISL_645829|20 ···················· ···················· ···················· ···················· ···················· ····················
R 6754 hCoV-19/England/ALDP-B1B8E1/2020|EPI_ISL_645830|20 ···················· ···················· ···················· ···················· ···················· ····················
R 6755 hCoV-19/England/ALDP-B1AC13/2020|EPI_ISL_645831|20 ···················· ···················· ···················· ···················· ···················· ····················
R 6756 hCoV-19/England/CAMC-B220D0/2020|EPI_ISL_645832|20 ···················· ···················· ···················· ···················· ···················· ····················
R 6757 hCoV-19/England/ALDP-B1F73B/2020|EPI_ISL_645833|20 ···················· ···················· ···················· ···················· ···················· ····················
R 6758 hCoV-19/England/ALDP-B1F9BD/2020|EPI_ISL_645845|20 ···················· ···················· ···················· ···················· ···················· ····················
R 6759 hCoV-19/England/ALDP-B1C0D9/2020|EPI_ISL_645846|20 ···················· ···················· ···················· ···················· ···················· ····················
R 6760 hCoV-19/England/ALDP-B1AE95/2020|EPI_ISL_645847|20 ···················· ···················· ···················· ···················· ···················· ····················
R 6761 hCoV-19/England/ALDP-B1F5D7/2020|EPI_ISL_645848|20 ···················· ···················· ···················· ···················· ···················· ····················
R 6762 hCoV-19/England/ALDP-B1ACD7/2020|EPI_ISL_645849|20 ···················· ···················· ···················· ···················· ···················· ····················
R 6763 hCoV-19/England/CAMC-B22252/2020|EPI_ISL_645850|20 ···················· ···················· ···················· ···················· ······T············· ····················
R 6764 hCoV-19/England/CAMC-B220FE/2020|EPI_ISL_645851|20 ···················· ···················· ···················· ···················· ······T············· ····················
R 6765 hCoV-19/England/CAMC-B22128/2020|EPI_ISL_645852|20 ···················· ···················· ···················· ···················· ···················· ····················
R 6766 hCoV-19/England/ALDP-B1AEE0/2020|EPI_ISL_645853|20 ···················· ···················· ···················· ···················· ···················· ····················
R 6767 hCoV-19/England/ALDP-B1F8ED/2020|EPI_ISL_645854|20 ···················· ···················· ···················· ···················· ···················· ····················
R 6768 hCoV-19/England/ALDP-B1DBC8/2020|EPI_ISL_645855|20 ···················· ···················· ···················· ···················· ···················· ····················

| | | | | | | | |
|---|---|---|---|---|---|---|---|
| R | 6769 hCoV-19/England/ALDP-B1DBD7/2020\|EPI_ISL_645856\|20 | ···················· | ···················· | ···················· | ···················· | ···················· | ···················· |
| R | 6770 hCoV-19/England/ALDP-B1AC8C/2020\|EPI_ISL_645857\|20 | ···················· | ···················· | ···················· | ···················· | ···················· | ···················· |
| R | 6771 hCoV-19/England/ALDP-B1D542/2020\|EPI_ISL_645858\|20 | ············T······· | ···················· | ···················· | ···················· | ···················· | ···················· |
| R | 6772 hCoV-19/England/ALDP-B1ACF5/2020\|EPI_ISL_645859\|20 | ···················· | ···················· | ···················· | ···················· | ···················· | ···················· |
| R | 6773 hCoV-19/England/ALDP-B1AE59/2020\|EPI_ISL_645860\|20 | ···················· | ···················· | ···················· | ···················· | ···················· | ···················· |
| R | 6774 hCoV-19/England/ALDP-B1DACB/2020\|EPI_ISL_645861\|20 | ···················· | ···················· | ···················· | ···················· | ···················· | ···················· |
| R | 6775 hCoV-19/England/ALDP-B1C09D/2020\|EPI_ISL_645862\|20 | ···················· | ···················· | ···················· | ···················· | ···················· | ···················· |
| R | 6776 hCoV-19/England/ALDP-B1C112/2020\|EPI_ISL_645863\|20 | ···················· | ···················· | ···················· | ···················· | ···················· | ···················· |
| R | 6777 hCoV-19/England/ALDP-B1ADC5/2020\|EPI_ISL_645864\|20 | ···················· | ···················· | ···················· | ···················· | ···················· | ···················· |
| R | 6778 hCoV-19/England/ALDP-B1D5AC/2020\|EPI_ISL_645865\|20 | ···················· | ······A············· | ···················· | ···················· | ···················· | ···················· |
| R | 6779 hCoV-19/England/CAMC-B2228F/2020\|EPI_ISL_645866\|20 | ···················· | ···················· | ···················· | ···················· | ···················· | ···················· |
| R | 6780 hCoV-19/England/ALDP-B1B8D2/2020\|EPI_ISL_645867\|20 | ···················· | ···················· | ···················· | ···················· | ···················· | ···················· |
| R | 6781 hCoV-19/England/CAMC-B22067/2020\|EPI_ISL_645868\|20 | ···················· | ···················· | ···················· | ···················· | ···················· | ···················· |
| R | 6782 hCoV-19/England/CAMC-B21D34/2020\|EPI_ISL_645869\|20 | ···················· | ·A·················· | ···················· | ···················· | ···················· | ···A················ |
| R | 6783 hCoV-19/England/ALDP-B1DAF8/2020\|EPI_ISL_645870\|20 | ···················· | ···················· | ···················· | ···················· | ···················· | ···················· |
| R | 6784 hCoV-19/England/ALDP-B1A2D5/2020\|EPI_ISL_645871\|20 | ···················· | ···················· | ···················· | ···················· | ···················· | ···················· |
| R | 6785 hCoV-19/England/ALDP-B1C0BB/2020\|EPI_ISL_645872\|20 | ···················· | ···················· | ···················· | ···················· | ···················· | ···················· |
| R | 6786 hCoV-19/England/ALDP-B1D4FA/2020\|EPI_ISL_645873\|20 | ···················· | ···················· | ···················· | ···················· | ···················· | ···················· |
| R | 6787 hCoV-19/England/ALDP-B1BCD6/2020\|EPI_ISL_645874\|20 | ···················· | ···················· | ···················· | ···················· | ···················· | ···················· |
| R | 6788 hCoV-19/England/ALDP-B1B298/2020\|EPI_ISL_645875\|20 | ···················· | ···················· | ···················· | ···················· | ···················· | ···················· |
| R | 6789 hCoV-19/England/ALDP-B1AC04/2020\|EPI_ISL_645876\|20 | ···················· | ···················· | ···················· | ···················· | ···················· | ···················· |
| R | 6790 hCoV-19/England/CAMC-B21FA7/2020\|EPI_ISL_645877\|20 | ···················· | ·A·······A·········· | ···················· | ···················· | ···················· | ···················· |
| R | 6791 hCoV-19/England/ALDP-B1A34B/2020\|EPI_ISL_645878\|20 | ···················· | ···················· | ···················· | ···················· | ···················· | ···················· |
| R | 6792 hCoV-19/England/ALDP-B1DB40/2020\|EPI_ISL_645879\|20 | ···················· | ···················· | ···················· | ···················· | ···················· | ···················· |
| R | 6793 hCoV-19/England/CAMC-B223C8/2020\|EPI_ISL_645880\|20 | ···················· | ···················· | ···················· | ···················· | ···················· | ···················· |
| R | 6794 hCoV-19/England/ALDP-B1B1F5/2020\|EPI_ISL_645881\|20 | ···················· | ···················· | ···················· | ···················· | ···················· | ···················· |
| R | 6795 hCoV-19/England/CAMC-B21C37/2020\|EPI_ISL_645882\|20 | ···················· | ·A·················· | ···················· | ···················· | ···················· | ···A················ |
| R | 6796 hCoV-19/England/ALDP-B1B896/2020\|EPI_ISL_645883\|20 | ···················· | ···················· | ···················· | ···················· | ···················· | ···················· |
| R | 6797 hCoV-19/England/ALDP-B1B8A5/2020\|EPI_ISL_645884\|20 | ···················· | ···················· | ···················· | ···················· | ···················· | ···················· |
| R | 6798 hCoV-19/England/CAMC-B22207/2020\|EPI_ISL_645885\|20 | ···················· | ···················· | ···················· | ···················· | ···················· | ···················· |
| R | 6799 hCoV-19/England/CAMC-B222F8/2020\|EPI_ISL_645886\|20 | ···················· | ···················· | ···················· | ···················· | ···················· | ···················· |
| R | 6800 hCoV-19/England/ALDP-B1F7A4/2020\|EPI_ISL_645887\|20 | ···················· | ···················· | ···················· | ···················· | ···················· | ···················· |
| R | 6801 hCoV-19/England/ALDP-B1F917/2020\|EPI_ISL_645888\|20 | ···················· | ···················· | ···················· | ···················· | ······T············· | ···················· |
| R | 6802 hCoV-19/England/ALDP-B1C130/2020\|EPI_ISL_645901\|20 | ···················· | ···················· | ···················· | ···················· | ···················· | ···················· |
| R | 6803 hCoV-19/England/CAMC-B225C2/2020\|EPI_ISL_645902\|20 | ···················· | ···················· | ···················· | ···················· | ···················· | ···················· |
| R | 6804 hCoV-19/England/ALDP-B1B7B7/2020\|EPI_ISL_645903\|20 | ···················· | ···················· | ···················· | ···················· | ···················· | ···················· |
| R | 6805 hCoV-19/England/CAMC-B22595/2020\|EPI_ISL_645905\|20 | ···················· | ·A·················· | ···················· | ···················· | ···················· | ···A················ |
| R | 6806 hCoV-19/England/ALDP-B1BD00/2020\|EPI_ISL_645906\|20 | ···················· | ···················· | ···················· | ···················· | ···················· | ···················· |
| R | 6807 hCoV-19/England/ALDP-B1A2C6/2020\|EPI_ISL_645907\|20 | ···················· | ···················· | ···················· | ···················· | ···················· | ···················· |
| R | 6808 hCoV-19/England/CAMC-B21E9B/2020\|EPI_ISL_645908\|20 | ···················· | ···················· | ···················· | ···················· | ···················· | ···················· |
| R | 6809 hCoV-19/England/ALDP-B1F7C2/2020\|EPI_ISL_645909\|20 | ···················· | ···················· | ···················· | ···················· | ···················· | ···················· |
| R | 6810 hCoV-19/England/ALDP-B1AE0E/2020\|EPI_ISL_645910\|20 | ···················· | ···················· | ···················· | ···················· | ···················· | ···················· |
| R | 6811 hCoV-19/England/ALDP-B1D533/2020\|EPI_ISL_645912\|20 | ···················· | ···················· | ···················· | ···················· | ···················· | ···················· |
| R | 6812 hCoV-19/England/ALDP-B1A299/2020\|EPI_ISL_645913\|20 | ···················· | ···················· | ···················· | ···················· | ···················· | ···················· |
| R | 6813 hCoV-19/England/ALDP-B1B78A/2020\|EPI_ISL_645914\|20 | ···················· | ···················· | ···················· | ···················· | ···················· | ···················· |
| R | 6814 hCoV-19/England/ALDP-B1AC31/2020\|EPI_ISL_645915\|20 | ···················· | ···················· | ···················· | ···················· | ···················· | ···················· |
| R | 6815 hCoV-19/England/CAMC-B22155/2020\|EPI_ISL_645916\|20 | ···················· | ···················· | ···················· | ···················· | ···················· | ···················· |
| R | 6816 hCoV-19/England/CAMC-B22410/2020\|EPI_ISL_645917\|20 | ···················· | ···················· | ···················· | ···················· | ···················· | ···················· |
| R | 6817 hCoV-19/England/CAMC-B2237D/2020\|EPI_ISL_645918\|20 | ···················· | ···················· | ···················· | ···················· | ···················· | ···················· |
| R | 6818 hCoV-19/England/ALDP-B1DB6E/2020\|EPI_ISL_645919\|20 | ···················· | ···················· | ···················· | ···················· | ···················· | ···················· |
| R | 6819 hCoV-19/England/ALDP-B1C0E8/2020\|EPI_ISL_645921\|20 | ···················· | ···················· | ···················· | ···················· | ···················· | ···················· |
| R | 6820 hCoV-19/Singapore/1172/2020\|EPI_ISL_648100\|2020-04 | ···················· | ···················· | ···················· | ···················· | ···················· | ···················· |
| R | 6821 hCoV-19/Singapore/1173/2020\|EPI_ISL_648101\|2020-04 | ···················· | ···················· | ···················· | ···················· | ···················· | ···················· |
| R | 6822 hCoV-19/Singapore/1174/2020\|EPI_ISL_648102\|2020-04 | ···················· | ···················· | ···················· | ···················· | ···················· | ···················· |
| R | 6823 hCoV-19/Singapore/1175/2020\|EPI_ISL_648103\|2020-04 | ···················· | ···················· | ···················· | ···················· | ···················· | ···················· |
| R | 6824 hCoV-19/Singapore/1176/2020\|EPI_ISL_648104\|2020-04 | ···················· | ···················· | ···················· | ···················· | ···················· | ···················· |
| R | 6825 hCoV-19/Singapore/1177/2020\|EPI_ISL_648105\|2020-04 | ···················· | ···················· | ···················· | ···················· | ···················· | ···················· |
| R | 6826 hCoV-19/Singapore/1178/2020\|EPI_ISL_648106\|2020-04 | ···················· | ···················· | ···················· | ···················· | ···················· | ···················· |
| R | 6827 hCoV-19/Singapore/1179/2020\|EPI_ISL_648107\|2020-04 | ···················· | ···················· | ···················· | ···················· | ···················· | ···················· |
| R | 6828 hCoV-19/Singapore/1180/2020\|EPI_ISL_648108\|2020-04 | ···················· | ···················· | ···················· | ···················· | ···················· | ···················· |
| R | 6829 hCoV-19/Singapore/1181/2020\|EPI_ISL_648109\|2020-04 | ···················· | ···················· | ···················· | ···················· | ···················· | ···················· |
| R | 6830 hCoV-19/Singapore/1182/2020\|EPI_ISL_648110\|2020-04 | ···················· | ···················· | ···················· | ···················· | ···················· | ···················· |
| R | 6831 hCoV-19/Singapore/1183/2020\|EPI_ISL_648111\|2020-04 | ···················· | ···················· | ···················· | ···················· | ···················· | ···················· |
| R | 6832 hCoV-19/Singapore/1184/2020\|EPI_ISL_648112\|2020-04 | ···················· | ···················· | ···················· | ···················· | ···················· | ···················· |
| R | 6833 hCoV-19/Singapore/1185/2020\|EPI_ISL_648113\|2020-04 | ···················· | ···················· | ···················· | ···················· | ···················· | ···················· |
| R | 6834 hCoV-19/Singapore/1186/2020\|EPI_ISL_648114\|2020-04 | ···················· | ···················· | ···················· | ···················· | ···················· | ···················· |
| R | 6835 hCoV-19/Singapore/1187/2020\|EPI_ISL_648115\|2020-04 | ···················· | ···················· | ···················· | ···················· | ···················· | ···················· |
| R | 6836 hCoV-19/Singapore/1188/2020\|EPI_ISL_648116\|2020-04 | ···················· | ···················· | ···················· | ···················· | ···················· | ···················· |
| R | 6837 hCoV-19/Singapore/1189/2020\|EPI_ISL_648117\|2020-04 | ···················· | ···················· | ···················· | ···················· | ···················· | ···················· |
| R | 6838 hCoV-19/Singapore/1190/2020\|EPI_ISL_648118\|2020-04 | ···················· | ···················· | ···················· | ···················· | ···················· | ···················· |
| R | 6839 hCoV-19/Singapore/1191/2020\|EPI_ISL_648119\|2020-04 | ···················· | ···················· | ···················· | ···················· | ···················· | ···················· |
| R | 6840 hCoV-19/Singapore/1192/2020\|EPI_ISL_648120\|2020-04 | ···················· | ···················· | ···················· | ···················· | ···················· | ···················· |
| R | 6841 hCoV-19/Singapore/1193/2020\|EPI_ISL_648121\|2020-04 | ···················· | ···················· | ···················· | ···················· | ···················· | ···················· |
| R | 6842 hCoV-19/Sweden/20-53278/2020\|EPI_ISL_648129\|2020-1 | ···················· | ···················· | ···················· | ···················· | ······T············· | ···················· |
| R | 6843 hCoV-19/Sweden/20-23091/2020\|EPI_ISL_648130\|2020-1 | ···················· | ···················· | ···················· | ···················· | ···················· | ···················· |
| R | 6844 hCoV-19/Sweden/20-23087/2020\|EPI_ISL_648131\|2020-0 | ···················· | ···················· | ···················· | ···················· | ···················· | ···················· |
| R | 6845 hCoV-19/Sweden/20-52765/2020\|EPI_ISL_648132\|2020-0 | ···················· | ·A·················· | ···················· | ···················· | ···················· | ···················· |
| R | 6846 hCoV-19/Sweden/20-52356/2020\|EPI_ISL_648133\|2020-0 | ···················· | ···················· | ···················· | ···················· | ···················· | ···················· |
| R | 6847 hCoV-19/Sweden/20-53092/2020\|EPI_ISL_648134\|2020-0 | ···················· | ···················· | ···················· | ···················· | ···················· | ···················· |
| R | 6848 hCoV-19/Sweden/20-53084/2020\|EPI_ISL_648135\|2020-1 | ···················· | ·A·················· | ···················· | ···················· | ···················· | ···················· |

R 6849 hCoV-19/Sweden/20-53086/2020|EPI_ISL_648136|2020-0 ···················· ····G··············· ···················· ···················· ···················· ····················
R 6850 hCoV-19/Sweden/20-53085/2020|EPI_ISL_648137|2020-1 ···················· ·A·················· ···················· ···················· ···················· ····················
R 6851 hCoV-19/Sweden/20-53091/2020|EPI_ISL_648138|2020-0 ···················· ···················· ···················· ···················· ···················· ····················
R 6852 hCoV-19/Sweden/20-53089/2020|EPI_ISL_648139|2020-0 ···················· ···················· ···················· ···················· ···················· ····················
R 6853 hCoV-19/Sweden/20-53056/2020|EPI_ISL_648140|2020-0 ···················· ···················· ···················· ···················· ···················· ····················
R 6854 hCoV-19/Sweden/20-53053/2020|EPI_ISL_648141|2020-0 ···················· ···················· ···················· ···················· ···················· ····················
R 6855 hCoV-19/Sweden/20-52695/2020|EPI_ISL_648142|2020-0 ···················· ····G··············· ···················· ···················· ···················· ····················
R 6856 hCoV-19/Sweden/20-53087/2020|EPI_ISL_648143|2020-0 ···················· ···················· ···················· ···················· ···················· ····················
R 6857 hCoV-19/Sweden/20-52813/2020|EPI_ISL_648144|2020-0 ···················· ···················· ···················· ···················· ···················· ····················
R 6858 hCoV-19/Sweden/20-53094/2020|EPI_ISL_648145|2020-0 ···················· ···················· ···················· ···················· ···················· ····················
R 6859 hCoV-19/Sweden/20-53079/2020|EPI_ISL_648146|2020-1 ···················· ···················· ···················· ···················· ···················· ····················
R 6860 hCoV-19/Sweden/20-53078/2020|EPI_ISL_648147|2020-1 ···················· ···················· ···················· ···················· ···················· ····················
R 6861 hCoV-19/Sweden/20-52881/2020|EPI_ISL_648148|2020-0 ···················· ···················· ···················· ···················· ···················· ····················
R 6862 hCoV-19/Sweden/20-53075/2020|EPI_ISL_648149|2020-1 ···················· ···················· ···················· ···················· ···················· ····················
R 6863 hCoV-19/Sweden/20-53076/2020|EPI_ISL_648150|2020-1 ···················· ···················· ···················· ···················· ···················· ····················
R 6864 hCoV-19/Sweden/20-53285/2020|EPI_ISL_648151|2020-1 ···················· ···················· ···················· ···················· ···················· ····················
R 6865 hCoV-19/Sweden/20-09857/2020|EPI_ISL_648152|2020-1 ···················· ···················· ···················· ···················· ···················· ····················
R 6866 hCoV-19/Sweden/20-53261/2020|EPI_ISL_648153|2020-1 ···················· ···················· ···················· ···················· ···················· ····················
R 6867 hCoV-19/Sweden/20-53257/2020|EPI_ISL_648154|2020-1 ···················· ···················· ···················· ···················· ···················· ····················
R 6868 hCoV-19/Sweden/20-53256/2020|EPI_ISL_648155|2020-1 ···················· ···················· ···················· ···················· ···················· ····················
R 6869 hCoV-19/Sweden/20-23682/2020|EPI_ISL_648156|2020-1 ···················· ···················· ···················· ···················· ···················· ····················
R 6870 hCoV-19/Sweden/20-53280/2020|EPI_ISL_648157|2020-1 ···················· ···················· ···················· ···················· ···················· ····················
R 6871 hCoV-19/Sweden/20-53279/2020|EPI_ISL_648158|2020-1 ···················· ···················· ···················· ···················· ···················· ····················
R 6872 hCoV-19/Sweden/20-53276/2020|EPI_ISL_648159|2020-1 ···················· ···················· ···················· ···················· ······T············· ····················
R 6873 hCoV-19/Sweden/20-53270/2020|EPI_ISL_648160|2020-1 ···················· ···················· ···················· ···················· ···················· ····················
R 6874 hCoV-19/Sweden/20-53269/2020|EPI_ISL_648161|2020-1 ···················· ···················· ···················· ···················· ···················· ····················
R 6875 hCoV-19/Sweden/20-53042/2020|EPI_ISL_648162|2020-1 ···················· ···················· ···················· ···················· ···················· ····················
R 6876 hCoV-19/Sweden/20-52820/2020|EPI_ISL_648163|2020-0 ···················· ···················· ···················· ···················· ···················· ····················
R 6877 hCoV-19/Sweden/20-53025/2020|EPI_ISL_648164|2020-0 ···················· ···················· ···················· ···················· ···················· ····················
R 6878 hCoV-19/Sweden/20-52728/2020|EPI_ISL_648165|2020-0 ···················· ···················· ···················· ···················· ···················· ····················
R 6879 hCoV-19/Sweden/20-53293/2020|EPI_ISL_648166|2020-1 ···················· ···················· ···················· ···················· ···················· ····················
R 6880 hCoV-19/Sweden/20-53191/2020|EPI_ISL_648167|2020-1 ···················· ···················· ···················· ···················· ···················· ····················
R 6881 hCoV-19/Sweden/20-53189/2020|EPI_ISL_648168|2020-1 ···················· ····A··············· ···················· ···················· ···················· ····················
R 6882 hCoV-19/Sweden/20-52742/2020|EPI_ISL_648169|2020-0 ···················· ···················· ···················· ···················· ···················· ····················
R 6883 hCoV-19/Sweden/20-53128/2020|EPI_ISL_648170|2020-0 ···················· ···················· ···················· ···················· ···················· ····················
R 6884 hCoV-19/Sweden/20-53127/2020|EPI_ISL_648171|2020-0 ···················· ·A·················· ···················· ···················· ···················· ····················
R 6885 hCoV-19/Sweden/20-53129/2020|EPI_ISL_648172|2020-0 ···················· ·A·················· ···················· ···················· ···················· ····················
R 6886 hCoV-19/Sweden/20-52391/2020|EPI_ISL_648173|2020-0 ···················· ···················· ···················· ···················· ···················· ····················
R 6887 hCoV-19/Sweden/20-53130/2020|EPI_ISL_648174|2020-0 ···················· ·A·················· ···················· ···················· ···················· ····················
R 6888 hCoV-19/Sweden/20-53131/2020|EPI_ISL_648175|2020-0 ···················· ···················· ···················· ···················· ···················· ····················
R 6889 hCoV-19/Sweden/20-53166/2020|EPI_ISL_648176|2020-1 ···················· ···················· ···················· ···················· ···················· ····················
R 6890 hCoV-19/Sweden/20-53165/2020|EPI_ISL_648177|2020-1 ···················· ···················· ···················· ···················· ······T············· ····················
R 6891 hCoV-19/Sweden/20-53169/2020|EPI_ISL_648178|2020-1 ···················· ···················· ···················· ···················· ···················· ····················
R 6892 hCoV-19/Sweden/20-53168/2020|EPI_ISL_648179|2020-1 ···················· ···················· ···················· ···················· ···················· ····················
R 6893 hCoV-19/Sweden/20-22151/2020|EPI_ISL_648180|2020-0 ···················· ···················· ···················· ···················· ···················· ····················
R 6894 hCoV-19/Sweden/20-08976/2020|EPI_ISL_648181|2020-0 ···················· ···················· ···················· ···················· ···················· ····················
R 6895 hCoV-19/Sweden/20-08977/2020|EPI_ISL_648182|2020-0 ···················· ···················· ···················· ···················· ···················· ····················
R 6896 hCoV-19/Sweden/20-08970/2020|EPI_ISL_648183|2020-0 ···················· ···················· ···················· ···················· ···················· ····················
R 6897 hCoV-19/Sweden/20-22144/2020|EPI_ISL_648184|2020-0 ···················· ···················· ···················· ···················· ···················· ····················
R 6898 hCoV-19/Sweden/20-23681/2020|EPI_ISL_648185|2020-1 ···················· ···················· ···················· ···················· ···················· ····················
R 6899 hCoV-19/Sweden/20-23688/2020|EPI_ISL_648186|2020-1 ···················· ·A·················· ···················· ···················· ···················· ····················
R 6900 hCoV-19/Sweden/20-23689/2020|EPI_ISL_648187|2020-1 ···················· ···················· ···················· ···················· ···················· ····················
R 6901 hCoV-19/Sweden/20-23684/2020|EPI_ISL_648188|2020-1 ···················· ···················· ···················· ···················· ···················· ····················
R 6902 hCoV-19/Sweden/20-23687/2020|EPI_ISL_648189|2020-1 ···················· ···················· ···················· ···················· ···················· ····················
R 6903 hCoV-19/Sweden/20-23686/2020|EPI_ISL_648190|2020-1 ···················· ···················· ···················· ···················· ···················· ····················
R 6904 hCoV-19/Sweden/20-23685/2020|EPI_ISL_648191|2020-1 ···················· ·A·················· ···················· ···················· ···················· ····················
R 6905 hCoV-19/Sweden/20-53195/2020|EPI_ISL_648192|2020-1 ···················· ···················· ···················· ···················· ···················· ····················
R 6906 hCoV-19/Sweden/20-52687/2020|EPI_ISL_648193|2020-0 ···················· ···················· ···················· ···················· ···················· ····················
R 6907 hCoV-19/Sweden/20-53102/2020|EPI_ISL_648194|2020-1 ···················· ···················· ···················· ···················· ···················· ····················
R 6908 hCoV-19/Sweden/20-53103/2020|EPI_ISL_648195|2020-0 ·····T·············· ···················· ···················· ···················· ···················· ····················
R 6909 hCoV-19/Sweden/20-53104/2020|EPI_ISL_648196|2020-0 ···················· ···················· ···················· ···················· ···················· ····················
R 6910 hCoV-19/Sweden/20-53105/2020|EPI_ISL_648197|2020-0 ···················· ····A··············· ···················· ···················· ···················· ····················
R 6911 hCoV-19/Sweden/20-52827/2020|EPI_ISL_648198|2020-0 ···················· ···················· ···················· ···················· ···················· ····················
R 6912 hCoV-19/Sweden/20-52828/2020|EPI_ISL_648199|2020-0 ···················· ···················· ···················· ···················· ···················· ····················
R 6913 hCoV-19/Russia/OMS-ORINFI-5368S/2020|EPI_ISL_63993 ···················· ···················· ···················· ···················· ···················· ····················
R 6914 hCoV-19/Russia/OMS-ORINFI-5753S/2020|EPI_ISL_63993 ···················· ···················· ···················· ···················· ···················· ····················
R 6915 hCoV-19/Russia/OMS-ORINFI-5774S/2020|EPI_ISL_63993 ···················· ···················· ···················· ···················· ···················· ····················
R 6916 hCoV-19/Russia/OMS-ORINFI-6212S/2020|EPI_ISL_63993 ···················· ···················· ···················· ···················· ···················· ····················
R 6917 hCoV-19/Russia/OMS-ORINFI-7871S/2020|EPI_ISL_63993 ···················· ···················· ···················· ···················· ···················· ····················
R 6918 hCoV-19/Russia/OMS-ORINFI-8293S/2020|EPI_ISL_63993 ···················· ···················· ···················· ···················· ···················· ····················
R 6919 hCoV-19/Russia/OMS-ORINFI-8296S/2020|EPI_ISL_63994 ···················· ···················· ···················· ···················· ···················· ····················
R 6920 hCoV-19/Russia/OMS-ORINFI-858S/2020|EPI_ISL_639941 ···················· ···················· ···················· ···················· ···················· ····················
R 6921 hCoV-19/Russia/OMS-ORINFI-848S/2020|EPI_ISL_639942 ···················· ···················· ···················· ···················· ···················· ····················
R 6922 hCoV-19/Russia/OMS-ORINFI-864S/2020|EPI_ISL_639943 ···················· ···················· ···················· ···················· ···················· ····················
R 6923 hCoV-19/Russia/OMS-ORINFI-865S/2020|EPI_ISL_639944 ···················· ···················· ···················· ···················· ···················· ····················
R 6924 hCoV-19/Russia/OMS-ORINFI-916S/2020|EPI_ISL_639945 ···················· ···················· ···················· ···················· ···················· ····················
R 6925 hCoV-19/Russia/OMS-ORINFI-9781S/2020|EPI_ISL_63994 ···················· ···················· ···················· ···················· ···················· ····················
R 6926 hCoV-19/Russia/OMS-ORINFI-9981S/2020|EPI_ISL_63994 ···················· ···················· ···················· ···················· ···················· ····················
R 6927 hCoV-19/Russia/SVE-RII-MH2828S/2020|EPI_ISL_639970 ···················· ···················· ···················· ···················· ···················· ····················
R 6928 hCoV-19/Russia/SVE-RII-MH2829S/2020|EPI_ISL_639971 ···················· ···················· ···················· ···················· ···················· ····················

R 6929 hCoV-19/Russia/SVE-RII-MH2830S/2020|EPI_ISL_639972 ···················· ···················· ···················· ···················· ···················· ····················
R 6930 hCoV-19/Russia/SVE-RII-MH2831S/2020|EPI_ISL_639973 ···················· ···················· ···················· ···················· ···················· ····················
R 6931 hCoV-19/France/ARA-NRC02715/2020|EPI_ISL_639974|20 ···················· ···················· ···················· ···················· ···················· ····················
R 6932 hCoV-19/France/PAC-NRC2718/2020|EPI_ISL_639975|202 ···················· ···················· ···················· ···················· ···················· ····················
R 6933 hCoV-19/France/ARA75790/2020|EPI_ISL_639976|2020-0 ···················· ···················· ···················· ···················· ···················· ····················
R 6934 hCoV-19/France/PAC-NRC02798/2020|EPI_ISL_639977|20 ···················· ···················· ···················· ···················· ···················· ····················
R 6935 hCoV-19/France/ARA74567/2020|EPI_ISL_639978|2020-0 ···················· ···················· ···················· ···················· ···················· ····················
R 6936 hCoV-19/France/ARA75377/2020|EPI_ISL_639979|2020-0 ···················· ···················· ···················· ···················· ···················· ····················
R 6937 hCoV-19/France/ARA76733/2020|EPI_ISL_639980|2020-0 ···················· ···················· ···················· ···················· ···················· ····················
R 6938 hCoV-19/France/ARA-NRC02745/2020|EPI_ISL_639981|20 ···················· ···················· ···················· ···················· ···················· ····················
R 6939 hCoV-19/France/ARA-NRC2813/2020|EPI_ISL_639983|202 ···················· ···················· ···················· ···················· ···················· ····················
R 6940 hCoV-19/France/ARA75924/2020|EPI_ISL_639984|2020-0 ···················· ···················· ···················· ···················· ···················· ····················
R 6941 hCoV-19/France/ARA75981/2020|EPI_ISL_639985|2020-0 ···················· ···················· ···················· ···················· ···················· ····················
R 6942 hCoV-19/France/ARA97420/2020|EPI_ISL_639986|2020-1 ···················· ···················· ···················· ···················· ···················· ····················
R 6943 hCoV-19/France/ARA-NRC02726/2020|EPI_ISL_639987|20 ···················· ···················· ···················· ···················· ···················· ····················
R 6944 hCoV-19/France/ARA-NRC2806/2020|EPI_ISL_639988|202 ···················· ···················· ···················· ···················· ···················· ····················
R 6945 hCoV-19/France/ARA97063/2020|EPI_ISL_639989|2020-1 ···················· ···················· ···················· ···················· ···················· ····················
R 6946 hCoV-19/France/ARA-NRC2802/2020|EPI_ISL_639990|202 ···················· ···················· ···················· ···················· ···················· ····················
R 6947 hCoV-19/France/ARA-NRC02706/2020|EPI_ISL_639991|20 ···················· ···················· ···················· ···················· ···················· ····················
R 6948 hCoV-19/France/ARA77081/2020|EPI_ISL_639992|2020-0 ···················· ···················· ···················· ···················· ···················· ····················
R 6949 hCoV-19/France/ARA78518/2020|EPI_ISL_639993|2020-0 ···················· ···················· ···················· ···················· ···················· ····················
R 6950 hCoV-19/France/ARA66285/2020|EPI_ISL_639994|2020-0 ···················· ···················· ···················· ···················· ···················· ····················
R 6951 hCoV-19/France/ARA78036/2020|EPI_ISL_639995|2020-0 ···················· ···················· ···················· ···················· ···················· ····················
R 6952 hCoV-19/France/PAC-NRC2803/2020|EPI_ISL_639996|202 ···················· ···················· ···················· ···················· ···················· ····················
R 6953 hCoV-19/France/PAC-NRC02800/2020|EPI_ISL_639997|20 ···················· ···················· ···················· ···················· ···················· ····················
R 6954 hCoV-19/France/ARA77080/2020|EPI_ISL_639998|2020-0 ···················· ···················· ···················· ···················· ···················· ····················
R 6955 hCoV-19/France/ARA68158/2020|EPI_ISL_639999|2020-0 ···················· ···················· ···················· ···················· ···················· ····················
R 6956 hCoV-19/Australia/VIC13231/2020|EPI_ISL_640931|202 ···················· ···················· ···················· ···················· ···················· ····················
R 6957 hCoV-19/Australia/VIC16073/2020|EPI_ISL_640932|202 ···················· ···················· ···················· ···················· ···················· ····················
R 6958 hCoV-19/Australia/VIC12691/2020|EPI_ISL_640934|202 ···················· ···················· ···················· ···················· ···················· ····················
R 6959 hCoV-19/Australia/VIC12636/2020|EPI_ISL_640935|202 ···················· ···················· ···················· ···················· ···················· ····················
R 6960 hCoV-19/Australia/VIC16558/2020|EPI_ISL_640936|202 ···················· ···················· ···················· ···················· ···················· ····················
R 6961 hCoV-19/Australia/VIC12289/2020|EPI_ISL_640938|202 ···················· ···················· ···················· ···················· ···················· ····················
R 6962 hCoV-19/Australia/VIC11828/2020|EPI_ISL_640939|202 ···················· ···················· ··K················· ···················· ···················· ····················
R 6963 hCoV-19/Australia/VIC16332/2020|EPI_ISL_640940|202 ···················· ···················· ···················· ···················· ···················· ····················
R 6964 hCoV-19/Australia/VIC16328/2020|EPI_ISL_640941|202 ···················· ···················· ···················· ···················· ···················· ····················
R 6965 hCoV-19/Australia/VIC16169/2020|EPI_ISL_640950|202 ···················· ···················· ···················· ···················· ···················· ····················
R 6966 hCoV-19/Australia/VIC12930/2020|EPI_ISL_640951|202 ···················· ···················· ···················· ···················· ···················· ····················
R 6968 hCoV-19/Australia/VIC13117/2020|EPI_ISL_640953|202 ···················· ···················· ···················· ···················· ···················· ····················
R 6969 hCoV-19/Australia/VIC16451/2020|EPI_ISL_640954|202 ···················· ···················· ···················· ···················· ···················· ····················
R 6970 hCoV-19/Australia/VIC12675/2020|EPI_ISL_640955|202 ···················· ···················· ···················· ···················· ···················· ····················
R 6971 hCoV-19/Australia/VIC13106/2020|EPI_ISL_640956|202 ···················· ···················· ···················· ···················· ···················· ····················
R 6972 hCoV-19/Australia/VIC12015/2020|EPI_ISL_640957|202 ···················· ···················· ···················· ···················· ···················· ····················
R 6973 hCoV-19/Australia/VIC12727/2020|EPI_ISL_640959|202 ···················· ···················· ···················· ···················· ···················· ····················
R 6974 hCoV-19/Australia/VIC12890/2020|EPI_ISL_640960|202 ···················· ···················· ···················· ···················· ···················· ····················
R 6975 hCoV-19/Australia/VIC13023/2020|EPI_ISL_640961|202 ···················· ···················· ···················· ···················· ···················· ····················
R 6976 hCoV-19/Australia/VIC16410/2020|EPI_ISL_640962|202 ···················· ···················· ···················· ···················· ···················· ····················
R 6978 hCoV-19/Australia/VIC16024/2020|EPI_ISL_640964|202 ···················· ···················· ···················· ···················· ···················· ····················
R 6979 hCoV-19/Australia/VIC16550/2020|EPI_ISL_640966|202 ···················· ···················· ···················· ···················· ···················· ····················
R 6980 hCoV-19/Australia/VIC16413/2020|EPI_ISL_640968|202 ···················· ···················· ···················· ···················· ···················· ····················
R 6981 hCoV-19/Australia/VIC16455/2020|EPI_ISL_640969|202 ···················· ···················· ···················· ···················· ···················· ····················
R 6982 hCoV-19/Australia/VIC12992/2020|EPI_ISL_640970|202 ···················· ···················· ···················· ···················· ···················· ····················
R 6984 hCoV-19/Australia/VIC16556/2020|EPI_ISL_640972|202 ···················· ···················· ···················· ···················· ···················· ····················
R 6985 hCoV-19/Australia/VIC15997/2020|EPI_ISL_640973|202 ···················· ···················· ···················· ···················· ···················· ····················
R 6987 hCoV-19/Australia/VIC12845/2020|EPI_ISL_640976|202 ···················· ···················· ···················· ···················· ···················· ····················
R 6989 hCoV-19/Australia/VIC16011/2020|EPI_ISL_640979|202 ···················· ···················· ···················· ···················· ···················· ····················
R 6990 hCoV-19/Australia/VIC16362/2020|EPI_ISL_640980|202 ···················· ···················· ···················· ···················· ···················· ····················
R 6991 hCoV-19/Australia/VIC12174/2020|EPI_ISL_640983|202 ···················· ···················· ···················· ···················· ···················· ····················
R 6992 hCoV-19/Australia/VIC12439/2020|EPI_ISL_640984|202 ···················· ···················· ···················· ···················· ···················· ····················
R 6993 hCoV-19/Australia/VIC15979/2020|EPI_ISL_640987|202 ···················· ···················· ···················· ···················· ···················· ····················
R 6994 hCoV-19/Australia/VIC16189/2020|EPI_ISL_640988|202 ···················· ···················· ···················· ···················· ···················· ····················
R 6995 hCoV-19/Australia/VIC16653/2020|EPI_ISL_640989|202 ···················· ···················· ···················· ···················· ··N················· ····················
R 6996 hCoV-19/Australia/VIC11612/2020|EPI_ISL_640991|202 ···················· ···················· ···················· ···················· ···················· ····················
R 6997 hCoV-19/Australia/VIC15885/2020|EPI_ISL_640992|202 ···················· ···················· ···················· ···················· ···················· ····················
R 6998 hCoV-19/Australia/VIC16132/2020|EPI_ISL_640993|202 ···················· ···················· ···················· ···················· ···················· ····················
R 6999 hCoV-19/England/QEUH-A8B9EA/2020|EPI_ISL_643227|20 ···················· ···················· ···················· ···················· ···················· ····················
R 7000 hCoV-19/England/QEUH-A8C503/2020|EPI_ISL_643229|20 ···················· ···················· ···················· ···················· ···················· ····················
R 7001 hCoV-19/England/QEUH-A8B5AA/2020|EPI_ISL_643230|20 ···················· ···················· ···················· ···················· ···················· ····················
R 7002 hCoV-19/England/QEUH-A8BA7E/2020|EPI_ISL_643232|20 ···················· ···················· ···················· ···················· ···················· ····················
R 7003 hCoV-19/England/QEUH-A8BCF0/2020|EPI_ISL_643233|20 ···················· ···················· ···················· ···················· ···················· ····················
R 7004 hCoV-19/England/QEUH-A8B55F/2020|EPI_ISL_643235|20 ···················· ···················· ···················· ···················· ···················· ····················
R 7005 hCoV-19/England/QEUH-A8C66A/2020|EPI_ISL_643237|20 ···················· ···················· ···················· ···················· ···················· ····················
R 7006 hCoV-19/England/QEUH-A8B63E/2020|EPI_ISL_643238|20 ···················· ···················· ···················· ···················· ···················· ····················
R 7007 hCoV-19/England/QEUH-A8BB11/2020|EPI_ISL_643241|20 ···················· ···················· ···················· ···················· ···················· ····················
R 7008 hCoV-19/England/QEUH-A8B513/2020|EPI_ISL_643242|20 ···················· ···················· ···················· ···················· ···················· ····················
R 7009 hCoV-19/England/QEUH-A8BE63/2020|EPI_ISL_643243|20 ···················· ···················· ···················· ···················· ···················· ····················
R 7010 hCoV-19/England/QEUH-A8C600/2020|EPI_ISL_643244|20 ···················· ···················· ···················· ···················· ···················· ····················
R 7011 hCoV-19/England/QEUH-A8B601/2020|EPI_ISL_643245|20 ···················· ·A·················· ···················· ···················· ···················· ····················
R 7012 hCoV-19/England/QEUH-A8C80A/2020|EPI_ISL_643246|20 ···················· ···················· ···················· ···················· ···················· ····················
R 7013 hCoV-19/England/QEUH-A8BC5A/2020|EPI_ISL_643247|20 ···················· ···················· ···················· ···················· ···················· ····················

| | | |
|---|---|---|
| R | 7014 hCoV-19/England/QEUH-A8B5E6/2020\|EPI_ISL_643248\|20 | ···················· ···················· ····················· ···················· ···················· ···················· |
| R | 7015 hCoV-19/England/QEUH-A8BC4B/2020\|EPI_ISL_643249\|20 | ···················· ···················· ····················· ···················· ···················· ···················· |
| R | 7016 hCoV-19/England/QEUH-A8BF51/2020\|EPI_ISL_643250\|20 | ···················· ···················· ····················· ···················· ···················· ···················· |
| R | 7017 hCoV-19/England/QEUH-A8B971/2020\|EPI_ISL_643251\|20 | ···················· ···················· ····················· ···················· ···················· ···················· |
| R | 7018 hCoV-19/England/QEUH-A8C2A2/2020\|EPI_ISL_643252\|20 | ···················· ···················· ····················· ···················· ···················· ···················· |
| R | 7019 hCoV-19/England/QEUH-A8C0E4/2020\|EPI_ISL_643253\|20 | ···················· ···················· ····················· ···················· ···················· ···················· |
| R | 7020 hCoV-19/England/QEUH-A8C63D/2020\|EPI_ISL_643256\|20 | ···················· ···················· ····················· ···················· ···················· ···················· |
| R | 7021 hCoV-19/England/QEUH-A8BECD/2020\|EPI_ISL_643257\|20 | ···················· ···················· ····················· ···················· ···················· ···················· |
| R | 7022 hCoV-19/England/QEUH-A8C934/2020\|EPI_ISL_643258\|20 | ···················· ···················· ····················· ···················· ···················· ···················· |
| R | 7023 hCoV-19/England/QEUH-A8B70E/2020\|EPI_ISL_643259\|20 | ···················· ···················· ····················· ···················· ···················· ···················· |
| R | 7024 hCoV-19/England/QEUH-A8B373/2020\|EPI_ISL_643260\|20 | ···················· ···················· ····················· ···················· ···················· ···················· |
| R | 7025 hCoV-19/England/QEUH-A8BEEB/2020\|EPI_ISL_643261\|20 | ···················· ···················· ····················· ···················· ···················· ···················· |
| R | 7026 hCoV-19/England/QEUH-A8C1F0/2020\|EPI_ISL_643262\|20 | ···················· ···················· ····················· ···················· ···················· ···················· |
| R | 7027 hCoV-19/England/QEUH-A8BCC3/2020\|EPI_ISL_643263\|20 | ···················· ···················· ····················· ···················· ···················· ···················· |
| R | 7028 hCoV-19/England/QEUH-A8B759/2020\|EPI_ISL_643264\|20 | ···················· ···················· ····················· ···················· ···················· ···················· |
| R | 7029 hCoV-19/England/QEUH-A8C688/2020\|EPI_ISL_643265\|20 | ···················· ···················· ····················· ···················· ···················· ···················· |
| R | 7030 hCoV-19/England/QEUH-A8C08A/2020\|EPI_ISL_643266\|20 | ···················· ···················· ····················· ···················· ···················· ···················· |
| R | 7031 hCoV-19/England/QEUH-A8C345/2020\|EPI_ISL_643267\|20 | ···················· ···················· ····················· ···················· ···················· ···················· |
| R | 7032 hCoV-19/England/QEUH-A8C98F/2020\|EPI_ISL_643268\|20 | ···················· ·A·················· ····················· ···················· ···················· ···················· |
| R | 7033 hCoV-19/England/QEUH-A8BF42/2020\|EPI_ISL_643269\|20 | ···················· ···················· ····················· ···················· ···················· ···················· |
| R | 7034 hCoV-19/England/QEUH-A8BE72/2020\|EPI_ISL_643270\|20 | ···················· ···················· ····················· ···················· ···················· ···················· |
| R | 7035 hCoV-19/England/QEUH-A8BCD2/2020\|EPI_ISL_643271\|20 | ···················· ···················· ····················· ···················· ···················· ···················· |
| R | 7036 hCoV-19/England/QEUH-A8C5C7/2020\|EPI_ISL_643272\|20 | ···················· ···················· ····················· ···················· ···················· ···················· |
| R | 7037 hCoV-19/England/QEUH-A8BBD5/2020\|EPI_ISL_643273\|20 | ···················· ···················· ····················· ···················· ···················· ···················· |
| R | 7038 hCoV-19/England/QEUH-A8B8B0/2020\|EPI_ISL_643274\|20 | ···················· ···················· ····················· ···················· ···················· ···················· |
| R | 7039 hCoV-19/England/QEUH-A8B8FC/2020\|EPI_ISL_643276\|20 | ···················· ···················· ····················· ···················· ···················· ···················· |
| R | 7040 hCoV-19/England/QEUH-A8BE18/2020\|EPI_ISL_643277\|20 | ···················· ···················· ····················· ···················· ···················· ···················· |
| R | 7041 hCoV-19/England/QEUH-A8B7D1/2020\|EPI_ISL_643278\|20 | ···················· ···················· ····················· ···················· ···················· ···················· |
| R | 7042 hCoV-19/England/QEUH-A8B4DA/2020\|EPI_ISL_643279\|20 | ···················· ···················· ····················· ···················· ···················· ···················· |
| R | 7043 hCoV-19/England/QEUH-A8C7A3/2020\|EPI_ISL_643280\|20 | ···················· ···················· ····················· ···················· ···················· ···················· |
| R | 7044 hCoV-19/England/QEUH-A8C64C/2020\|EPI_ISL_643281\|20 | ···················· ···················· ····················· ···················· ···················· ···················· |
| R | 7045 hCoV-19/England/QEUH-A8B58C/2020\|EPI_ISL_643282\|20 | ···················· ···················· ····················· ···················· ···················· ···················· |
| R | 7046 hCoV-19/England/QEUH-A8C318/2020\|EPI_ISL_643283\|20 | ···················· ···················· ····················· ···················· ···················· ···················· |
| R | 7047 hCoV-19/England/QEUH-A8C3FA/2020\|EPI_ISL_643284\|20 | ···················· ···················· ····················· ···················· ···················· ···················· |
| R | 7048 hCoV-19/England/QEUH-A8C284/2020\|EPI_ISL_643285\|20 | ···················· ···················· ····················· ···················· ···················· ···················· |
| R | 7049 hCoV-19/England/QEUH-A8C196/2020\|EPI_ISL_643288\|20 | ···················· ···················· ····················· ···················· ···················· ···················· |
| R | 7050 hCoV-19/England/QEUH-A8BE54/2020\|EPI_ISL_643292\|20 | ···················· ···················· ····················· ···················· ···················· ···················· |
| R | 7051 hCoV-19/England/QEUH-A8C309/2020\|EPI_ISL_643293\|20 | ···················· ···················· ····················· ···················· ···················· ···················· |
| R | 7052 hCoV-19/England/QEUH-A8B319/2020\|EPI_ISL_643294\|20 | ···················· ···················· ····················· ···················· ···················· ···················· |
| R | 7053 hCoV-19/England/QEUH-A8B838/2020\|EPI_ISL_643295\|20 | ···················· ···················· ····················· ···················· ···················· ···················· |
| R | 7054 hCoV-19/England/QEUH-A8BAC9/2020\|EPI_ISL_643296\|20 | ···················· ···················· ····················· ···················· ···················· ···················· |
| R | 7055 hCoV-19/England/QEUH-A8C9F8/2020\|EPI_ISL_643297\|20 | ···················· ···················· ····················· ···················· ···················· ···················· |
| R | 7056 hCoV-19/England/QEUH-A8C943/2020\|EPI_ISL_643298\|20 | ···················· ···················· ····················· ···················· ···················· ···················· |
| R | 7057 hCoV-19/England/QEUH-A8B908/2020\|EPI_ISL_643299\|20 | ···················· ···················· ····················· ···················· ···················· ···················· |
| R | 7058 hCoV-19/France/PAC-MEPHI-2116/2020\|EPI_ISL_644501\| | ···················· ···················· ····················· ···················· ···················· ···················· |
| R | 7059 hCoV-19/France/PAC-MEPHI-2118/2020\|EPI_ISL_644503\| | ···················· ···················· ····················· ···················· ···················· ···················· |
| R | 7060 hCoV-19/France/PAC-MEPHI-2128/2020\|EPI_ISL_644504\| | ···················· ···················· ····················· ···················· ···················· ···················· |
| R | 7061 hCoV-19/France/PAC-MEPHI-2132/2020\|EPI_ISL_644505\| | ···················· ···················· ····················· ···················· ···················· ···················· |
| R | 7062 hCoV-19/France/PAC-MEPHI-2135/2020\|EPI_ISL_644506\| | ···················· ···················· ····················· ···················· ···················· ···················· |
| R | 7063 hCoV-19/France/PAC-MEPHI-2136/2020\|EPI_ISL_644507\| | ···················· ···················· ····················· ···················· ···················· ···················· |
| R | 7064 hCoV-19/France/PAC-MEPHI-2139/2020\|EPI_ISL_644508\| | ···················· ···················· ····················· ···················· ···················· ···················· |
| R | 7065 hCoV-19/France/PAC-MEPHI-2140/2020\|EPI_ISL_644509\| | ···················· ···················· ····················· ···················· ···················· ···················· |
| R | 7066 hCoV-19/France/PAC-MEPHI-2144/2020\|EPI_ISL_644510\| | ···················· ···················· ····················· ···················· ···················· ···················· |
| R | 7067 hCoV-19/France/PAC-MEPHI-2147/2020\|EPI_ISL_644511\| | ···················· ···················· ····················· ···················· ···················· ···················· |
| R | 7068 hCoV-19/France/PAC-MEPHI-2148/2020\|EPI_ISL_644512\| | ···················· ···················· ····················· ···················· ···················· ···················· |
| R | 7069 hCoV-19/France/PAC-MEPHI-2151/2020\|EPI_ISL_644513\| | ···················· ···················· ····················· ···················· ···················· ···················· |
| R | 7070 hCoV-19/France/PAC-MEPHI-2402/2020\|EPI_ISL_644558\| | ···················· ···················· ····················· ···················· ···················· ···················· |
| R | 7071 hCoV-19/France/PAC-MEPHI-2410/2020\|EPI_ISL_644559\| | ···················· ···················· ····················· ···················· ···················· ···················· |
| R | 7072 hCoV-19/France/PAC-MEPHI-2411/2020\|EPI_ISL_644560\| | ···················· ···················· ····················· ···················· ···················· ···················· |
| R | 7073 hCoV-19/France/PAC-MEPHI-2413/2020\|EPI_ISL_644561\| | ···················· ···················· ····················· ···················· ···················· ···················· |
| R | 7074 hCoV-19/France/PAC-MEPHI-2414/2020\|EPI_ISL_644562\| | ···················· ···················· ····················· ···················· ···················· ···················· |
| R | 7075 hCoV-19/France/PAC-MEPHI-2416/2020\|EPI_ISL_644563\| | ···················· ···················· ····················· ···················· ···················· ···················· |
| R | 7076 hCoV-19/France/PAC-MEPHI-2417/2020\|EPI_ISL_644564\| | ···················· ···················· ····················· ···················· ···················· ···················· |
| R | 7077 hCoV-19/Serbia/Kosovska Mitrovica-05221607/2020\|EP | ···················· ···················· ····················· ···················· ···················· ···················· |
| R | 7078 hCoV-19/Serbia/Kosovska Mitrovica-08372107/2020\|EP | ···················· ···················· ····················· ···················· ···················· ···················· |
| R | 7079 hCoV-19/Serbia/Kosovska Mitrovica-09872307/2020\|EP | ···················· ···················· ····················· ···················· ···················· ···················· |
| R | 7080 hCoV-19/Serbia/Kraljevo-00040707/2020\|EPI_ISL_6445 | ···················· ···················· ····················· ···················· ···················· ···················· |
| R | 7081 hCoV-19/Serbia/Kraljevo-00152307/2020\|EPI_ISL_6445 | ···················· ···················· ····················· ···················· ···················· ···················· |
| R | 7082 hCoV-19/Serbia/Kraljevo-00160707/2020\|EPI_ISL_6445 | ···················· ···················· ····················· ···················· ···················· ···················· |
| R | 7083 hCoV-19/Serbia/Kraljevo-00221607/2020\|EPI_ISL_6445 | ···················· ···················· ····················· ···················· ···················· ···················· |
| R | 7084 hCoV-19/Serbia/Kraljevo-00521205/2020\|EPI_ISL_6445 | ···················· ···················· ····················· ···················· ···················· ···················· |
| R | 7085 hCoV-19/Serbia/Kraljevo-00571207/2020\|EPI_ISL_6445 | ···················· ···················· ····················· ···················· ···················· ···················· |
| R | 7086 hCoV-19/Serbia/Kraljevo-02872507/2020\|EPI_ISL_6445 | ···················· ···················· ····················· ···················· ···················· ···················· |
| R | 7087 hCoV-19/Serbia/Kraljevo-02912507/2020\|EPI_ISL_6445 | ···················· ···················· ····················· ···················· ···················· ···················· |
| R | 7088 hCoV-19/Serbia/Kraljevo-03082507/2020\|EPI_ISL_6445 | ···················· ···················· ····················· ···················· ···················· ···················· |
| R | 7089 hCoV-19/Serbia/Kraljevo-03102507/2020\|EPI_ISL_6445 | ···················· ···················· ····················· ···················· ···················· ···················· |
| R | 7090 hCoV-19/Serbia/Kraljevo-03592507/2020\|EPI_ISL_6445 | ···················· ···················· ····················· ···················· ···················· ···················· |
| R | 7091 hCoV-19/Serbia/Novi Pazar-00391007/2020\|EPI_ISL_64 | ···················· ···················· ····················· ···················· ···················· ···················· |
| R | 7092 hCoV-19/Serbia/Novi Pazar-17611405/2020\|EPI_ISL_64 | ···················· ···················· ····················· ···················· ···················· ···················· |
| R | 7093 hCoV-19/Serbia/Novi Pazar-17641405/2020\|EPI_ISL_64 | ···················· ···················· ····················· ···················· ···················· ···················· |

| | | | | | | | |
|---|---|---|---|---|---|---|---|
| R | 7094 hCoV-19/Serbia/Uzice-00200704/2020\|EPI_ISL_644582\| | ···················· | ···················· | ···················· | ···················· | ···················· | ···················· |
| R | 7095 hCoV-19/Serbia/Uzice-01450604/2020\|EPI_ISL_644583\| | ···················· | ···················· | ···················· | ···················· | ···················· | ···················· |
| R | 7096 hCoV-19/USA/NM-NMDOH-2020327015/2020\|EPI_ISL_64458 | ···················· | ···················· | ···················· | ···················· | ···················· | ···················· |
| R | 7097 hCoV-19/USA/NM-NMDOH-2020328211/2020\|EPI_ISL_64458 | ···················· | ···················· | ···················· | ···················· | ···················· | ···················· |
| R | 7098 hCoV-19/USA/NM-NMDOH-2020328277/2020\|EPI_ISL_64458 | ···················· | ···················· | ···················· | ···················· | ···················· | ···················· |
| R | 7099 hCoV-19/USA/NM-NMDOH-2020328328/2020\|EPI_ISL_64459 | ···················· | ···················· | ···················· | ···················· | ···················· | ···················· |
| R | 7100 hCoV-19/USA/NM-NMDOH-2020328744/2020\|EPI_ISL_64459 | ···T················ | ···················· | ···················· | ···················· | ···················· | ···················· |
| R | 7101 hCoV-19/USA/NM-NMDOH-2020328756/2020\|EPI_ISL_64459 | ···T················ | ···················· | ···················· | ···················· | ···················· | ···················· |
| R | 7102 hCoV-19/USA/NM-NMDOH-2020328776/2020\|EPI_ISL_64459 | ···················· | ···················· | ···················· | ···················· | ···················· | ···················· |
| R | 7103 hCoV-19/USA/NM-NMDOH-2020328986/2020\|EPI_ISL_64459 | ···················· | ·A·················· | ···················· | ···················· | ···················· | ···················· |
| R | 7104 hCoV-19/USA/NM-NMDOH-2020329046/2020\|EPI_ISL_64459 | ···················· | ···················· | ···················· | ···················· | ···················· | ···················· |
| R | 7105 hCoV-19/USA/NM-NMDOH-2020329048/2020\|EPI_ISL_64459 | ···················· | ···················· | ···················· | ···················· | ···················· | ···················· |
| R | 7106 hCoV-19/USA/NM-NMDOH-2020329049/2020\|EPI_ISL_64459 | ···················· | ···················· | ···················· | ···················· | ···················· | ···················· |
| R | 7107 hCoV-19/USA/NM-NMDOH-2020329076/2020\|EPI_ISL_64459 | ···················· | ···················· | ···················· | ···················· | ···················· | ···················· |
| R | 7108 hCoV-19/USA/NM-NMDOH-2020329922/2020\|EPI_ISL_64459 | ···················· | ···················· | ···················· | ···················· | ···················· | ···················· |
| R | 7109 hCoV-19/England/ALDP-B1B571/2020\|EPI_ISL_645801\|20 | ···················· | ···················· | ···················· | ···················· | ···················· | ···················· |
| R | 7110 hCoV-19/England/ALDP-B1C516/2020\|EPI_ISL_645803\|20 | ···················· | ···················· | ···················· | ···················· | ···················· | ···················· |
| R | 7111 hCoV-19/England/ALDP-B1A712/2020\|EPI_ISL_645804\|20 | ···················· | ···················· | ···················· | ···················· | ···················· | ···················· |
| R | 7112 hCoV-19/England/ALDP-B1B31D/2020\|EPI_ISL_645805\|20 | ···················· | ···················· | ···················· | ···················· | ···················· | ···················· |
| R | 7113 hCoV-19/England/ALDP-B1B438/2020\|EPI_ISL_645806\|20 | ···················· | ···················· | ···················· | ···················· | ···················· | ···················· |
| R | 7114 hCoV-19/England/ALDP-B1CF36/2020\|EPI_ISL_645807\|20 | ···················· | ···················· | ···················· | ···················· | ···················· | ···················· |
| R | 7115 hCoV-19/England/ALDP-B1B4CF/2020\|EPI_ISL_645808\|20 | ···················· | ···················· | ···················· | ···················· | ···················· | ···················· |
| R | 7116 hCoV-19/England/ALDP-B1BBAC/2020\|EPI_ISL_645810\|20 | ···················· | ···················· | ···················· | ···················· | ···················· | ···················· |
| R | 7117 hCoV-19/England/ALDP-B1BA36/2020\|EPI_ISL_645811\|20 | ···················· | ···················· | ···················· | ···················· | ···················· | ···················· |
| R | 7118 hCoV-19/England/ALDP-B1B27A/2020\|EPI_ISL_645812\|20 | ···················· | ···················· | ···················· | ···················· | ···················· | ···················· |
| R | 7119 hCoV-19/England/ALDP-B1B201/2020\|EPI_ISL_645813\|20 | ···················· | ···················· | ···················· | ···················· | ···················· | ···················· |
| R | 7120 hCoV-19/England/ALDP-B1B9C0/2020\|EPI_ISL_645814\|20 | ···················· | ···················· | ···················· | ···················· | ···················· | ···················· |
| R | 7121 hCoV-19/England/ALDP-B1BBE8/2020\|EPI_ISL_645815\|20 | ···················· | ···················· | ···················· | ···················· | ···················· | ···················· |
| R | 7122 hCoV-19/England/ALDP-B1B4A1/2020\|EPI_ISL_645816\|20 | ···················· | ···················· | ···················· | ···················· | ···················· | ···················· |
| R | 7123 hCoV-19/England/ALDP-B1B4DE/2020\|EPI_ISL_645817\|20 | ···················· | ···················· | ···················· | ···················· | ···················· | ···················· |
| R | 7124 hCoV-19/England/ALDP-B1B33B/2020\|EPI_ISL_645818\|20 | ···················· | ···················· | ···················· | ···················· | ···················· | ···················· |
| R | 7125 hCoV-19/England/ALDP-B1A1BA/2020\|EPI_ISL_645819\|20 | ···················· | ···················· | ···················· | ···················· | ···················· | ···················· |
| R | 7126 hCoV-19/England/ALDP-B1C534/2020\|EPI_ISL_645820\|20 | ···················· | ···················· | ···················· | ···················· | ···················· | ···················· |
| R | 7127 hCoV-19/England/ALDP-B1CF18/2020\|EPI_ISL_645821\|20 | ···················· | ···················· | ···················· | ···················· | ···················· | ···················· |
| R | 7128 hCoV-19/England/ALDP-B1AFB0/2020\|EPI_ISL_645822\|20 | ···················· | ···················· | ···················· | ···················· | ···················· | ···················· |
| R | 7129 hCoV-19/England/ALDP-B1AD01/2020\|EPI_ISL_645834\|20 | ···················· | ···················· | ···················· | ···················· | ···················· | ···················· |
| R | 7130 hCoV-19/England/ALDP-B1D506/2020\|EPI_ISL_645835\|20 | ···················· | ···················· | ···················· | ···················· | ···················· | ···················· |
| R | 7131 hCoV-19/England/ALDP-B1AE77/2020\|EPI_ISL_645837\|20 | ···················· | ···················· | ···················· | ···················· | ···················· | ···················· |
| R | 7132 hCoV-19/England/CAMC-B22261/2020\|EPI_ISL_645838\|20 | ···················· | ···················· | ···················· | ···················· | ······T············· | ···················· |
| R | 7133 hCoV-19/England/CAMC-B22322/2020\|EPI_ISL_645840\|20 | ·T·················· | ···················· | ···················· | ···················· | ···················· | ···················· |
| R | 7134 hCoV-19/England/ALDP-B1A41B/2020\|EPI_ISL_645841\|20 | ···················· | ···················· | ···················· | ···················· | ···················· | ···················· |
| R | 7135 hCoV-19/England/ALDP-B1F65C/2020\|EPI_ISL_645842\|20 | ···················· | ···················· | ···················· | ···················· | ···················· | ···················· |
| R | 7136 hCoV-19/England/ALDP-B1F7E0/2020\|EPI_ISL_645843\|20 | ···················· | ···················· | ···················· | ···················· | ···················· | ···················· |
| R | 7137 hCoV-19/England/ALDP-B1C051/2020\|EPI_ISL_645844\|20 | ···················· | ···················· | ···················· | ···················· | ···················· | ···················· |
| R | 7138 hCoV-19/England/ALDP-B1B289/2020\|EPI_ISL_645889\|20 | ···················· | ···················· | ···················· | ···················· | ···················· | ···················· |
| R | 7139 hCoV-19/England/ALDP-B1F847/2020\|EPI_ISL_645890\|20 | ···················· | ···················· | ···················· | ···················· | ······T············· | ···················· |
| R | 7140 hCoV-19/England/ALDP-B1A3F0/2020\|EPI_ISL_645891\|20 | ···················· | ···················· | ···················· | ···················· | ···················· | ···················· |
| R | 7141 hCoV-19/England/ALDP-B1C14F/2020\|EPI_ISL_645892\|20 | ···················· | ···················· | ···················· | ···················· | ···················· | ···················· |
| R | 7142 hCoV-19/England/ALDP-B1AC9B/2020\|EPI_ISL_645893\|20 | ···················· | ···················· | ···················· | ···················· | ···················· | ···················· |
| R | 7143 hCoV-19/England/ALDP-B1AD4D/2020\|EPI_ISL_645894\|20 | ···················· | ···················· | ···················· | ···················· | ···················· | ···················· |
| R | 7144 hCoV-19/England/ALDP-B1C07F/2020\|EPI_ISL_645895\|20 | ···················· | ···················· | ···················· | ···················· | ···················· | ···················· |
| R | 7145 hCoV-19/England/CAMC-B2260B/2020\|EPI_ISL_645896\|20 | ···················· | ···················· | ···················· | ···················· | ···················· | ···················· |
| R | 7146 hCoV-19/England/CAMC-B222DA/2020\|EPI_ISL_645897\|20 | ···················· | ···················· | ···················· | ···················· | ···················· | ···················· |
| R | 7147 hCoV-19/England/ALDP-B1AF29/2020\|EPI_ISL_645898\|20 | ···················· | ···················· | ···················· | ···················· | ···················· | ···················· |
| R | 7148 hCoV-19/England/CAMC-B21D43/2020\|EPI_ISL_645899\|20 | ···················· | ···················· | ···················· | ···················· | ···················· | ···················· |
| R | 7149 hCoV-19/Singapore/136nan/2020\|EPI_ISL_648080\|2020- | ···················· | ···················· | ···················· | ···················· | ···················· | ···················· |
| R | 7150 hCoV-19/Singapore/1155/2020\|EPI_ISL_648081\|2020-03 | ···················· | ···················· | ···················· | ···················· | ···················· | ···················· |
| R | 7151 hCoV-19/Singapore/1156/2020\|EPI_ISL_648082\|2020-03 | ···················· | ···················· | ···················· | ···················· | ···················· | ···················· |
| R | 7152 hCoV-19/Singapore/1157/2020\|EPI_ISL_648083\|2020-03 | ···················· | ···················· | ···················· | ···················· | ···················· | ···················· |
| R | 7153 hCoV-19/Singapore/1158/2020\|EPI_ISL_648084\|2020-03 | ···················· | ···················· | ···················· | ···················· | ···················· | ···················· |
| R | 7154 hCoV-19/Singapore/1159/2020\|EPI_ISL_648085\|2020-03 | ···················· | ···················· | ···················· | ···················· | ···················· | ···················· |
| R | 7155 hCoV-19/Singapore/907nan/2020\|EPI_ISL_648086\|2020- | ···················· | ···················· | ···················· | ···················· | ···················· | ···················· |
| R | 7156 hCoV-19/Singapore/1160/2020\|EPI_ISL_648087\|2020-04 | ···················· | ···················· | ···················· | ···················· | ···················· | ···················· |
| R | 7157 hCoV-19/Singapore/1161/2020\|EPI_ISL_648088\|2020-04 | ···················· | ···················· | ···················· | ···················· | ···················· | ···················· |
| R | 7158 hCoV-19/Singapore/1162/2020\|EPI_ISL_648089\|2020-04 | ···················· | ···················· | ···················· | ···················· | ···················· | ···················· |
| R | 7159 hCoV-19/Singapore/560nan/2020\|EPI_ISL_648090\|2020- | ···················· | ···················· | ···················· | ···················· | ···················· | ···················· |
| R | 7160 hCoV-19/Singapore/1163/2020\|EPI_ISL_648091\|2020-04 | ···················· | ···················· | ···················· | ···················· | ···················· | ···················· |
| R | 7161 hCoV-19/Singapore/1164/2020\|EPI_ISL_648092\|2020-04 | ···················· | ···················· | ···················· | ···················· | ···················· | ···················· |
| R | 7162 hCoV-19/Singapore/1165/2020\|EPI_ISL_648093\|2020-04 | ···················· | ···················· | ···················· | ···················· | ···················· | ···················· |
| R | 7163 hCoV-19/Singapore/1166/2020\|EPI_ISL_648094\|2020-04 | ···················· | ···················· | ···················· | ···················· | ···················· | ···················· |
| R | 7164 hCoV-19/Singapore/1167/2020\|EPI_ISL_648095\|2020-04 | ···················· | ···················· | ···················· | ···················· | ···················· | ···················· |
| R | 7165 hCoV-19/Singapore/1168/2020\|EPI_ISL_648096\|2020-04 | ···················· | ···················· | ···················· | ···················· | ···················· | ···················· |
| R | 7166 hCoV-19/Singapore/1169/2020\|EPI_ISL_648097\|2020-04 | ···················· | ···················· | ···················· | ···················· | ···················· | ···················· |
| R | 7167 hCoV-19/Singapore/1170/2020\|EPI_ISL_648098\|2020-04 | ···················· | ···················· | ···················· | ···················· | ···················· | ···················· |
| R | 7168 hCoV-19/Singapore/1171/2020\|EPI_ISL_648099\|2020-04 | ···················· | ···················· | ···················· | ···················· | ···················· | ···················· |
| R | 7169 hCoV-19/Ireland/MN-NVRL-20IRL50633/2020\|EPI_ISL_63 | ···················· | ···················· | ···················· | ···················· | ···················· | ···················· |
| R | 7170 hCoV-19/Ireland/MN-NVRL-20IRL50451/2020\|EPI_ISL_63 | ···················· | ···················· | ···················· | ···················· | ···················· | ···················· |
| R | 7171 hCoV-19/Ireland/MN-NVRL-20IRL51554/2020\|EPI_ISL_63 | ···················· | ···················· | ···················· | ···················· | ···················· | ···················· |
| R | 7172 hCoV-19/Ireland/MN-NVRL-20IRL51503/2020\|EPI_ISL_63 | ···················· | ···················· | ···················· | ···················· | ···················· | ···················· |
| R | 7173 hCoV-19/Ireland/CN-NVRL-20IRL82829/2020\|EPI_ISL_63 | ···················· | ···················· | ···················· | ···················· | ···················· | ···················· |

| | | | | | | | | |
|---|---|---|---|---|---|---|---|---|
| R | 7174 hCoV-19/Russia/OMS-ORINFI-9916S/2020\|EPI_ISL_63992 | ···················· | ···················· | ···················· | ···················· | ···················· | ···················· | |
| R | 7175 hCoV-19/Russia/OMS-ORINFI-5311S/2020\|EPI_ISL_63992 | ···················· | ···················· | ···················· | ···················· | ···················· | ···················· | |
| R | 7176 hCoV-19/Russia/OMS-ORINFI-5312S/2020\|EPI_ISL_63992 | ···················· | ···················· | ···················· | ···················· | ···················· | ···················· | |
| R | 7177 hCoV-19/Russia/OMS-ORINFI-6390S/2020\|EPI_ISL_63993 | ···················· | ···················· | ···················· | ···················· | ···················· | ···················· | |
| R | 7178 hCoV-19/Russia/OMS-ORINFI-6680S/2020\|EPI_ISL_63993 | ···················· | ···················· | ···················· | ···················· | ···················· | ···················· | |
| R | 7179 hCoV-19/Russia/OMS-ORINFI-7131S/2020\|EPI_ISL_63993 | ···················· | ···················· | ···················· | ···················· | ···················· | ···················· | |
| R | 7180 hCoV-19/Russia/OMS-ORINFI-7139S/2020\|EPI_ISL_63993 | ···················· | ···················· | ···················· | ···················· | ···················· | ···················· | |
| R | 7181 hCoV-19/Russia/PER-RII-MH1647S/2020\|EPI_ISL_639949 | ···················· | ···················· | ···················· | ···················· | ···················· | ···················· | |
| R | 7182 hCoV-19/Russia/MOS-RII-MH2800S/2020\|EPI_ISL_639951 | ···················· | ···················· | ···················· | ···················· | ···················· | ···················· | |
| R | 7183 hCoV-19/Russia/MOS-RII-MH2802S/2020\|EPI_ISL_639952 | ···················· | ···················· | ···················· | ···················· | ···················· | ···················· | |
| R | 7184 hCoV-19/Russia/MOS-RII-MH2804S/2020\|EPI_ISL_639953 | ···················· | ···················· | ···················· | ···················· | ···················· | ···················· | |
| R | 7185 hCoV-19/Russia/MOS-RII-MH2805S/2020\|EPI_ISL_639954 | ···················· | ···················· | ···················· | ···················· | ···················· | ···················· | |
| R | 7186 hCoV-19/Russia/MOS-RII-MH2806S/2020\|EPI_ISL_639955 | ···················· | ···················· | ···················· | ···················· | ···················· | ···················· | |
| R | 7187 hCoV-19/Russia/MOS-RII-MH2807S/2020\|EPI_ISL_639956 | ···················· | ···················· | ···················· | ···················· | ···················· | ···················· | |
| R | 7188 hCoV-19/Russia/MOS-RII-MH2808S/2020\|EPI_ISL_639957 | ···················· | ···················· | ···················· | ···················· | ···················· | ···················· | |
| R | 7189 hCoV-19/Russia/NVS-RII-MH2809S/2020\|EPI_ISL_639958 | ···················· | ···················· | ···················· | ···················· | ···················· | ···················· | |
| R | 7190 hCoV-19/Russia/NVS-RII-MH2810S/2020\|EPI_ISL_639959 | ···················· | ···················· | ···················· | ···················· | ···················· | ···················· | |
| R | 7191 hCoV-19/Russia/ORE-RII-MH2814S/2020\|EPI_ISL_639961 | ···················· | ···················· | ···················· | ···················· | ···················· | ···················· | |
| R | 7192 hCoV-19/Russia/PER-RII-MH2816S/2020\|EPI_ISL_639963 | ···················· | ···················· | ···················· | ···················· | ···················· | ···················· | |
| R | 7193 hCoV-19/Russia/PER-RII-MH2818S/2020\|EPI_ISL_639964 | ···················· | ···················· | ···················· | ···················· | ···················· | ···················· | |
| R | 7194 hCoV-19/Russia/ROS-RII-MH2821S/2020\|EPI_ISL_639966 | ···················· | ···················· | ···················· | ···················· | ···················· | ···················· | |
| R | 7195 hCoV-19/Russia/RYA-RII-MH2823S/2020\|EPI_ISL_639967 | ···················· | ···················· | ·········T·········· | ···················· | ···················· | ···················· | |
| R | 7196 hCoV-19/Russia/RYA-RII-MH2824S/2020\|EPI_ISL_639968 | ···················· | ···················· | ···················· | ···················· | ···················· | ···················· | |
| R | 7197 hCoV-19/Russia/SVE-RII-MH2824S/2020\|EPI_ISL_639969 | ···················· | ···················· | ···················· | ···················· | ···················· | ···················· | |
| R | 7198 hCoV-19/Australia/VIC12516/2020\|EPI_ISL_640911\|202 | ···················· | ···················· | ···················· | ···················· | ···················· | ···················· | |
| R | 7199 hCoV-19/Australia/VIC12371/2020\|EPI_ISL_640913\|202 | ···················· | ···················· | ···················· | ···················· | NNN················· | ···················· | 1 |
| R | 7201 hCoV-19/Australia/VIC15996/2020\|EPI_ISL_640915\|202 | ···················· | ···················· | ···················· | ···················· | ···················· | ···················· | |
| R | 7202 hCoV-19/Australia/VIC11998/2020\|EPI_ISL_640916\|202 | ···················· | ···················· | ···················· | ···················· | ···················· | ···················· | |
| R | 7204 hCoV-19/Australia/VIC12906/2020\|EPI_ISL_640918\|202 | ···················· | ···················· | ···················· | ···················· | ···················· | ···················· | |
| R | 7205 hCoV-19/Australia/VIC13110/2020\|EPI_ISL_640919\|202 | ···················· | ···················· | ···················· | ···················· | NNN················· | ···················· | 1 |
| R | 7206 hCoV-19/Australia/VIC16025/2020\|EPI_ISL_640921\|202 | ···················· | ···················· | ···················· | ···················· | ···················· | ···················· | |
| R | 7208 hCoV-19/Australia/VIC12028/2020\|EPI_ISL_640923\|202 | ···················· | ···················· | ···················· | ···················· | ···················· | ···················· | |
| R | 7209 hCoV-19/Australia/VIC11975/2020\|EPI_ISL_640924\|202 | ···················· | ···················· | ···················· | ···················· | ···················· | ···················· | |
| R | 7210 hCoV-19/Australia/VIC12087/2020\|EPI_ISL_640925\|202 | ···················· | ···················· | ···················· | ···················· | ···················· | ···················· | |
| R | 7211 hCoV-19/Australia/VIC16048/2020\|EPI_ISL_640926\|202 | ···················· | ···················· | ···················· | ···················· | ···················· | ···················· | |
| R | 7212 hCoV-19/Australia/VIC16076/2020\|EPI_ISL_640928\|202 | ···················· | ···················· | ···················· | ···················· | ···················· | ···················· | |
| R | 7213 hCoV-19/Australia/VIC12905/2020\|EPI_ISL_640943\|202 | ···················· | ······A············· | ···················· | ···················· | ···················· | ···················· | |
| R | 7214 hCoV-19/Australia/VIC16544/2020\|EPI_ISL_640944\|202 | ···················· | ···················· | ···················· | ···················· | ···················· | ···················· | |
| R | 7215 hCoV-19/Australia/VIC16026/2020\|EPI_ISL_640945\|202 | ···················· | ···················· | ···················· | ···················· | ···················· | ···················· | |
| R | 7217 hCoV-19/Australia/VIC13086/2020\|EPI_ISL_640948\|202 | ···················· | ···················· | ···················· | ···················· | ···················· | ···················· | |
| R | 7218 hCoV-19/Australia/VIC12222/2020\|EPI_ISL_640994\|202 | ···················· | ···················· | ···················· | ···················· | ···················· | ···················· | |
| R | 7219 hCoV-19/Australia/VIC16438/2020\|EPI_ISL_640995\|202 | ···················· | ···················· | ···················· | ···················· | ···················· | ···················· | |
| R | 7220 hCoV-19/Australia/VIC12821/2020\|EPI_ISL_640996\|202 | ···················· | ···················· | ···················· | ···················· | ···················· | ···················· | |
| R | 7222 hCoV-19/Australia/VIC12091/2020\|EPI_ISL_640999\|202 | ···················· | ···················· | ···················· | ···················· | ···················· | ···················· | |
| R | 7223 hCoV-19/England/QEUH-A8B8DE/2020\|EPI_ISL_643100\|20 | ···················· | ···················· | ···················· | ···················· | ···················· | ···················· | |
| R | 7224 hCoV-19/England/QEUH-A8C48E/2020\|EPI_ISL_643101\|20 | ···················· | ···················· | ···················· | ···················· | ···················· | ···················· | |
| R | 7225 hCoV-19/England/QEUH-A8BBF3/2020\|EPI_ISL_643102\|20 | ···················· | ···················· | ···················· | ···················· | ···················· | ···················· | |
| R | 7226 hCoV-19/England/QEUH-A8C5E5/2020\|EPI_ISL_643104\|20 | ···················· | ···················· | ···················· | ···················· | ···················· | ···················· | |
| R | 7227 hCoV-19/England/QEUH-A8BA14/2020\|EPI_ISL_643170\|20 | ···················· | ···················· | ···················· | ···················· | ···················· | ···················· | |
| R | 7228 hCoV-19/England/QEUH-A8C7C1/2020\|EPI_ISL_643171\|20 | ···················· | ···················· | ···················· | ···················· | ···················· | ···················· | |
| R | 7229 hCoV-19/England/QEUH-A8B4AD/2020\|EPI_ISL_643172\|20 | ···················· | ···················· | ···················· | ···················· | ···················· | ···················· | |
| R | 7230 hCoV-19/England/QEUH-A8B67A/2020\|EPI_ISL_643173\|20 | ···················· | ···················· | ···················· | ···················· | ···················· | ···················· | |
| R | 7231 hCoV-19/England/QEUH-A8BA50/2020\|EPI_ISL_643174\|20 | ···················· | ···················· | ···················· | ···················· | ···················· | ···················· | |
| R | 7232 hCoV-19/England/QEUH-A8C925/2020\|EPI_ISL_643175\|20 | ···················· | ···················· | ···················· | ···················· | ···················· | ···················· | |
| R | 7233 hCoV-19/England/QEUH-A8C275/2020\|EPI_ISL_643179\|20 | ···················· | ···················· | ···················· | ···················· | ···················· | ···················· | |
| R | 7234 hCoV-19/England/QEUH-A8BEBE/2020\|EPI_ISL_643190\|20 | ···················· | ···················· | ···················· | ···················· | ···················· | ···················· | |
| R | 7235 hCoV-19/England/QEUH-A8C5D6/2020\|EPI_ISL_643191\|20 | ···················· | ···················· | ···················· | ···················· | ···················· | ···················· | |
| R | 7236 hCoV-19/England/QEUH-A8BB20/2020\|EPI_ISL_643192\|20 | ···················· | ···················· | ···················· | ···················· | ···················· | ···················· | |
| R | 7237 hCoV-19/England/QEUH-A8B883/2020\|EPI_ISL_643193\|20 | ···················· | ···················· | ···················· | ···················· | ···················· | ···················· | |
| R | 7238 hCoV-19/England/QEUH-A8C06C/2020\|EPI_ISL_643194\|20 | ···················· | ·A·················· | ···················· | ···················· | ···················· | ···A················ | |
| R | 7239 hCoV-19/England/QEUH-A8C406/2020\|EPI_ISL_643195\|20 | ···················· | ···················· | ···················· | ···················· | ···················· | ···················· | |
| R | 7240 hCoV-19/England/QEUH-A8B3BF/2020\|EPI_ISL_643196\|20 | ···················· | ···················· | ···················· | ···················· | ···················· | ···················· | |
| R | 7241 hCoV-19/England/QEUH-A8C11E/2020\|EPI_ISL_643197\|20 | ···················· | ···················· | ···················· | ···················· | ···················· | ···················· | |
| R | 7242 hCoV-19/England/QEUH-A8C49D/2020\|EPI_ISL_643198\|20 | ···················· | ···················· | ···················· | ···················· | ···················· | ···················· | |
| R | 7243 hCoV-19/England/QEUH-A8C354/2020\|EPI_ISL_643199\|20 | ···················· | ···················· | ···················· | ···················· | ···················· | ···················· | |
| R | 7244 hCoV-19/France/PAC-MEPHI-1682/2020\|EPI_ISL_644400\| | ···················· | ···················· | ···················· | ···················· | ···················· | ···················· | |
| R | 7245 hCoV-19/France/PAC-MEPHI-1731/2020\|EPI_ISL_644401\| | ···················· | ···················· | ···················· | ···················· | ···················· | ···················· | |
| R | 7246 hCoV-19/France/PAC-MEPHI-0143/2020\|EPI_ISL_644403\| | ···················· | ···················· | ···················· | ···················· | ···················· | ···················· | |
| R | 7247 hCoV-19/France/PAC-MEPHI-0312/2020\|EPI_ISL_644404\| | ···················· | ··········A········· | ···················· | ···················· | ···················· | ···················· | |
| R | 7248 hCoV-19/France/PAC-MEPHI-0650/2020\|EPI_ISL_644405\| | ···················· | ···················· | ···················· | ···················· | ···················· | ···················· | |
| R | 7249 hCoV-19/France/PAC-MEPHI-0651/2020\|EPI_ISL_644406\| | ···················· | ···················· | ···················· | ···················· | ···················· | ···················· | |
| R | 7250 hCoV-19/France/PAC-MEPHI-0652/2020\|EPI_ISL_644407\| | ···················· | ···················· | ···················· | ···················· | ···················· | ···················· | |
| R | 7251 hCoV-19/France/PAC-MEPHI-0749/2020\|EPI_ISL_644408\| | ···················· | ···················· | ···················· | ···················· | ···················· | ···················· | |
| R | 7252 hCoV-19/France/PAC-MEPHI-0752/2020\|EPI_ISL_644409\| | ···················· | ···················· | ···················· | ···················· | ···················· | ···················· | |
| R | 7253 hCoV-19/France/PAC-MEPHI-0762/2020\|EPI_ISL_644410\| | ···················· | ···················· | ···················· | ···················· | ···················· | ···················· | |
| R | 7254 hCoV-19/France/PAC-MEPHI-1613/2020\|EPI_ISL_644411\| | ···················· | ···················· | ···················· | ···················· | ···················· | ···················· | |
| R | 7255 hCoV-19/France/PAC-MEPHI-1614/2020\|EPI_ISL_644412\| | ···················· | ···················· | ···················· | ···················· | ···················· | ···················· | |
| R | 7256 hCoV-19/France/PAC-MEPHI-1667/2020\|EPI_ISL_644413\| | ···················· | ···················· | ···················· | ···················· | ···················· | ···················· | |
| R | 7257 hCoV-19/France/PAC-MEPHI-1669/2020\|EPI_ISL_644414\| | ···················· | ···················· | ···················· | ···················· | ···················· | ···················· | |
| R | 7258 hCoV-19/France/PAC-MEPHI-1670/2020\|EPI_ISL_644415\| | ···················· | ···················· | ···················· | ···················· | ···················· | ···················· | |

| | | | | | | | | |
|---|---|---|---|---|---|---|---|---|
| R | 7259 | hCoV-19/France/PAC-MEPHI-1671/2020\|EPI_ISL_644416\| | ···················· | ···················· | ···················· | ···················· | ···················· | ···················· |
| R | 7260 | hCoV-19/France/PAC-MEPHI-1673/2020\|EPI_ISL_644417\| | ···················· | ···················· | ···················· | ···················· | ···················· | ···················· |
| R | 7261 | hCoV-19/France/PAC-MEPHI-1674/2020\|EPI_ISL_644418\| | ···················· | ···················· | ···················· | ···················· | ···················· | ···················· |
| R | 7262 | hCoV-19/France/PAC-MEPHI-1675/2020\|EPI_ISL_644419\| | ···················· | ···················· | ···················· | ···················· | ···················· | ···················· |
| R | 7263 | hCoV-19/France/PAC-MEPHI-1676/2020\|EPI_ISL_644420\| | ···················· | ···················· | ···················· | ···················· | ···················· | ···················· |
| R | 7264 | hCoV-19/France/PAC-MEPHI-1677/2020\|EPI_ISL_644421\| | ···················· | ···················· | ···················· | ···················· | ···················· | ···················· |
| R | 7265 | hCoV-19/France/PAC-MEPHI-1678/2020\|EPI_ISL_644422\| | ···················· | ···················· | ···················· | ···················· | ···················· | ···················· |
| R | 7266 | hCoV-19/France/PAC-MEPHI-1679/2020\|EPI_ISL_644423\| | ···················· | ···················· | ···················· | ···················· | ···················· | ···················· |
| R | 7267 | hCoV-19/France/PAC-MEPHI-1681/2020\|EPI_ISL_644424\| | ···················· | ···················· | ···················· | ···················· | ···················· | ···················· |
| R | 7268 | hCoV-19/France/PAC-MEPHI-1697/2020\|EPI_ISL_644425\| | ···················· | ···················· | ···················· | ···················· | ···················· | ···················· |
| R | 7269 | hCoV-19/France/PAC-MEPHI-1702/2020\|EPI_ISL_644426\| | ···················· | ···················· | ···················· | ···················· | ···················· | ···················· |
| R | 7270 | hCoV-19/France/PAC-MEPHI-1703/2020\|EPI_ISL_644427\| | ···················· | ···················· | ···················· | ···················· | ···················· | ···················· |
| R | 7271 | hCoV-19/France/PAC-MEPHI-1716/2020\|EPI_ISL_644428\| | ···················· | ···················· | ···················· | ···················· | ···················· | ···················· |
| R | 7272 | hCoV-19/France/PAC-MEPHI-1718/2020\|EPI_ISL_644429\| | ···················· | ···················· | ···················· | ···················· | ···················· | ···················· |
| R | 7273 | hCoV-19/France/PAC-MEPHI-1719/2020\|EPI_ISL_644430\| | ···················· | ···················· | ···················· | ···················· | ···················· | ···················· |
| R | 7274 | hCoV-19/France/PAC-MEPHI-1720/2020\|EPI_ISL_644431\| | ···················· | ···················· | ···················· | ···················· | ···················· | ···················· |
| R | 7275 | hCoV-19/France/PAC-MEPHI-1723/2020\|EPI_ISL_644432\| | ···················· | ···················· | ···················· | ···················· | ···················· | ···················· |
| R | 7276 | hCoV-19/France/PAC-MEPHI-1724/2020\|EPI_ISL_644433\| | ···················· | ···················· | ···················· | ···················· | ···················· | ···················· |
| R | 7277 | hCoV-19/France/PAC-MEPHI-1729/2020\|EPI_ISL_644434\| | ···················· | ···················· | ···················· | ···················· | ···················· | ···················· |
| R | 7278 | hCoV-19/France/PAC-MEPHI-1730/2020\|EPI_ISL_644435\| | ···················· | ···················· | ···················· | ···················· | ···················· | ···················· |
| R | 7279 | hCoV-19/France/PAC-MEPHI-1733/2020\|EPI_ISL_644436\| | ···················· | ···················· | ···················· | ···················· | ···················· | ···················· |
| R | 7280 | hCoV-19/England/ALDP-B1A74F/2020\|EPI_ISL_645700\|20 | ···················· | ···················· | ···················· | ···················· | ···················· | ···················· |
| R | 7281 | hCoV-19/England/ALDP-B1B140/2020\|EPI_ISL_645701\|20 | ···················· | ···················· | ···················· | ···················· | ···················· | ···················· |
| R | 7282 | hCoV-19/England/ALDP-B1ABAD/2020\|EPI_ISL_645702\|20 | ···················· | ···················· | ···················· | ···················· | ···················· | ···················· |
| R | 7283 | hCoV-19/England/ALDP-B1BF37/2020\|EPI_ISL_645703\|20 | ···················· | ···················· | ···················· | ···················· | ···················· | ···················· |
| R | 7284 | hCoV-19/England/ALDP-B1CD4B/2020\|EPI_ISL_645704\|20 | ···················· | ···················· | ···················· | ···················· | ···················· | ···················· |
| R | 7285 | hCoV-19/England/ALDP-B1BB7F/2020\|EPI_ISL_645706\|20 | ···················· | ···················· | ···················· | ···················· | ···················· | ···················· |
| R | 7286 | hCoV-19/England/ALDP-B1BB8E/2020\|EPI_ISL_645707\|20 | ···················· | ···················· | ···················· | ···················· | ···················· | ···················· |
| R | 7287 | hCoV-19/England/ALDP-B1B544/2020\|EPI_ISL_645708\|20 | ···················· | ···················· | ···················· | ···················· | ·····T·············· | ···················· |
| R | 7288 | hCoV-19/England/ALDP-B1C3C1/2020\|EPI_ISL_645709\|20 | ···················· | ···················· | ···················· | ···················· | ···················· | ···················· |
| R | 7289 | hCoV-19/England/ALDP-B1AB9E/2020\|EPI_ISL_645710\|20 | ···················· | ···················· | ···················· | ···················· | ···················· | ···················· |
| R | 7290 | hCoV-19/England/ALDP-B1ABBC/2020\|EPI_ISL_645711\|20 | ···················· | ···················· | ···················· | ···················· | ···················· | ···················· |
| R | 7291 | hCoV-19/England/ALDP-B1CEDF/2020\|EPI_ISL_645712\|20 | ···················· | ···················· | ···················· | ···················· | ···················· | ···················· |
| R | 7292 | hCoV-19/England/ALDP-B1CEA2/2020\|EPI_ISL_645713\|20 | ···················· | ···················· | ···················· | ···················· | ···················· | ···················· |
| R | 7293 | hCoV-19/England/ALDP-B1E1D4/2020\|EPI_ISL_645714\|20 | ···················· | ···················· | ···················· | ···················· | ···················· | ···················· |
| R | 7294 | hCoV-19/England/ALDP-B1CD78/2020\|EPI_ISL_645715\|20 | ···················· | ···················· | ···················· | ···················· | ···················· | ···················· |
| R | 7295 | hCoV-19/England/ALDP-B1B052/2020\|EPI_ISL_645716\|20 | ···················· | ·A·················· | ···················· | ···················· | ···················· | ···················· |
| R | 7296 | hCoV-19/England/ALDP-B1CF27/2020\|EPI_ISL_645717\|20 | ···················· | ···················· | ···················· | ···················· | ···················· | ···················· |
| R | 7297 | hCoV-19/England/ALDP-B1C570/2020\|EPI_ISL_645718\|20 | ···················· | ···················· | ···················· | ···················· | ···················· | ···················· |
| R | 7298 | hCoV-19/England/ALDP-B1B9A2/2020\|EPI_ISL_645719\|20 | ···················· | ···················· | ···················· | ···················· | ···················· | ···················· |
| R | 7299 | hCoV-19/England/ALDP-B1B131/2020\|EPI_ISL_645720\|20 | ···················· | ···················· | ···················· | ···················· | ···················· | ···················· |
| R | 7300 | hCoV-19/England/ALDP-B1BF0A/2020\|EPI_ISL_645721\|20 | ···················· | ···················· | ···················· | ···················· | ···················· | ···················· |
| R | 7301 | hCoV-19/England/ALDP-B1C4EC/2020\|EPI_ISL_645722\|20 | ···················· | ·A·················· | ···················· | ···················· | ···················· | ···················· |
| R | 7302 | hCoV-19/England/ALDP-B1B948/2020\|EPI_ISL_645723\|20 | ···················· | ···················· | ···················· | ···················· | ···················· | ···················· |
| R | 7303 | hCoV-19/England/ALDP-B1CDC3/2020\|EPI_ISL_645724\|20 | ···················· | ···················· | ···················· | ···················· | ···················· | ···················· |
| R | 7304 | hCoV-19/England/ALDP-B1E1C5/2020\|EPI_ISL_645725\|20 | ···················· | ···················· | ···················· | ···················· | ···················· | ···················· |
| R | 7305 | hCoV-19/England/ALDP-B1BC12/2020\|EPI_ISL_645726\|20 | ···················· | ···················· | ···················· | ···················· | ···················· | ···················· |
| R | 7306 | hCoV-19/England/ALDP-B1BA45/2020\|EPI_ISL_645727\|20 | ···················· | ···················· | ···················· | ···················· | ···················· | ···················· |
| R | 7307 | hCoV-19/England/ALDP-B1B59F/2020\|EPI_ISL_645728\|20 | ···················· | ···················· | ···················· | ···················· | ···················· | ···················· |
| R | 7308 | hCoV-19/England/ALDP-B1CE75/2020\|EPI_ISL_645729\|20 | ···················· | ···················· | ···················· | ···················· | ···················· | ···················· |
| R | 7309 | hCoV-19/England/ALDP-B1A1C9/2020\|EPI_ISL_645730\|20 | ···················· | ···················· | ···················· | ···················· | ···················· | ···················· |
| R | 7310 | hCoV-19/England/ALDP-B1A0BD/2020\|EPI_ISL_645731\|20 | ···················· | ···················· | ···················· | ···················· | ···················· | ···················· |
| R | 7311 | hCoV-19/England/ALDP-B1BF73/2020\|EPI_ISL_645732\|20 | ···················· | ···················· | ···················· | ···················· | ···················· | ···················· |
| R | 7312 | hCoV-19/England/ALDP-B1A651/2020\|EPI_ISL_645733\|20 | ···················· | ···················· | ···················· | ···················· | ···················· | ···················· |
| R | 7313 | hCoV-19/England/ALDP-B1B359/2020\|EPI_ISL_645734\|20 | ···················· | ···················· | ···················· | ···················· | ···················· | ···················· |
| R | 7314 | hCoV-19/England/ALDP-B1A0DB/2020\|EPI_ISL_645735\|20 | ···················· | ···················· | ···················· | ···················· | ···················· | ···················· |
| R | 7315 | hCoV-19/England/ALDP-B1B30E/2020\|EPI_ISL_645736\|20 | ···················· | ···················· | ···················· | ···················· | ···················· | ··A················· |
| R | 7316 | hCoV-19/England/ALDP-B1BA81/2020\|EPI_ISL_645737\|20 | ···················· | ···················· | ···················· | ···················· | ···················· | ···················· |
| R | 7317 | hCoV-19/England/ALDP-B1CDB4/2020\|EPI_ISL_645738\|20 | ···················· | ···················· | ···················· | ···················· | ···················· | ···················· |
| R | 7318 | hCoV-19/England/ALDP-B1CEB1/2020\|EPI_ISL_645740\|20 | ···················· | ···················· | ···················· | ···················· | ···················· | ···················· |
| R | 7319 | hCoV-19/England/ALDP-B1B0CB/2020\|EPI_ISL_645741\|20 | ···················· | ···················· | ···················· | ···················· | ···················· | ···················· |
| R | 7320 | hCoV-19/England/ALDP-B1B09E/2020\|EPI_ISL_645742\|20 | ···················· | ···················· | ···················· | ···················· | ···················· | ···················· |
| R | 7321 | hCoV-19/England/ALDP-B1CE1B/2020\|EPI_ISL_645743\|20 | ···················· | ···················· | ···················· | ···················· | ···················· | ···················· |
| R | 7322 | hCoV-19/England/ALDP-B19F57/2020\|EPI_ISL_645744\|20 | ···················· | ···················· | ···················· | ···················· | ·····T·············· | ···················· |
| R | 7323 | hCoV-19/England/ALDP-B1B08F/2020\|EPI_ISL_645745\|20 | ···················· | ···················· | ···················· | ···················· | ···················· | ···················· |
| R | 7324 | hCoV-19/England/ALDP-B1C385/2020\|EPI_ISL_645746\|20 | ···················· | ···················· | ···················· | ···················· | ···················· | ···················· |
| R | 7325 | hCoV-19/England/ALDP-B1CD5A/2020\|EPI_ISL_645747\|20 | ···················· | ···················· | ···················· | ···················· | ···················· | ···················· |
| R | 7326 | hCoV-19/England/ALDP-B1E277/2020\|EPI_ISL_645748\|20 | ···················· | ···················· | ···················· | ···················· | ···················· | ···················· |
| R | 7327 | hCoV-19/England/ALDP-B1B386/2020\|EPI_ISL_645749\|20 | ···················· | ···················· | ···················· | ···················· | ···················· | ···················· |
| R | 7328 | hCoV-19/England/ALDP-B1A624/2020\|EPI_ISL_645750\|20 | ···················· | ···················· | ···················· | ···················· | ·····T·············· | ···················· |
| R | 7329 | hCoV-19/England/ALDP-B1B456/2020\|EPI_ISL_645751\|20 | ···················· | ···················· | ···················· | ···················· | ···················· | ···················· |
| R | 7330 | hCoV-19/England/ALDP-B1C3A3/2020\|EPI_ISL_645752\|20 | ···················· | ···················· | ···················· | ···················· | ···················· | ···················· |
| R | 7331 | hCoV-19/England/ALDP-B1C015/2020\|EPI_ISL_645753\|20 | ···················· | ···················· | ···················· | ···················· | ···················· | ···················· |
| R | 7332 | hCoV-19/England/ALDP-B1AF74/2020\|EPI_ISL_645754\|20 | ···················· | ···················· | ···················· | ···················· | ···················· | ···················· |
| R | 7333 | hCoV-19/England/ALDP-B1B22F/2020\|EPI_ISL_645755\|20 | ···················· | ···················· | ···················· | ···················· | ···················· | ···················· |
| R | 7334 | hCoV-19/England/ALDP-B1CE57/2020\|EPI_ISL_645756\|20 | ···················· | ···················· | ···················· | ···················· | ···················· | ···················· |
| R | 7335 | hCoV-19/England/ALDP-B1A721/2020\|EPI_ISL_645757\|20 | ···················· | ···················· | ···················· | ···················· | ···················· | ···················· |
| R | 7336 | hCoV-19/England/ALDP-B1A141/2020\|EPI_ISL_645758\|20 | ···················· | ···················· | ···················· | ···················· | ···················· | ···················· |
| R | 7337 | hCoV-19/England/ALDP-B1C59E/2020\|EPI_ISL_645759\|20 | ···················· | ···················· | ···················· | ···················· | ···················· | ···················· |
| R | 7338 | hCoV-19/England/ALDP-B19F0C/2020\|EPI_ISL_645760\|20 | ···················· | ···················· | ···················· | ···················· | ·····T·············· | ···················· |

| | | | | | | | | |
|---|---|---|---|---|---|---|---|---|
| R | 7339 | hCoV-19/England/ALDP-B1BADC/2020\|EPI_ISL_645761\|20 | ···················· | ···················· | ···················· | ···················· | ···················· | ···················· |
| R | 7340 | hCoV-19/England/ALDP-B1B18C/2020\|EPI_ISL_645762\|20 | ···················· | ·A·················· | ···················· | ···················· | ···················· | ···················· |
| R | 7341 | hCoV-19/England/ALDP-B1ABDA/2020\|EPI_ISL_645764\|20 | ···················· | ···················· | ···················· | ···················· | ···················· | ···················· |
| R | 7342 | hCoV-19/England/ALDP-B1BFEC/2020\|EPI_ISL_645765\|20 | ···················· | ···················· | ···················· | ···················· | ···················· | ···················· |
| R | 7343 | hCoV-19/England/ALDP-B1B535/2020\|EPI_ISL_645767\|20 | ···················· | ···················· | ···················· | ···················· | ···················· | ···················· |
| R | 7344 | hCoV-19/England/ALDP-B1CF09/2020\|EPI_ISL_645800\|20 | ···················· | ···················· | ···················· | ···················· | ···················· | ···················· |
| R | 7345 | hCoV-19/USA/IA-CDC-4054/2020\|EPI_ISL_648000\|2020-0 | ···················· | ···················· | ···················· | ···················· | ···················· | ···················· |
| R | 7346 | hCoV-19/USA/IA-CDC-4055/2020\|EPI_ISL_648001\|2020-0 | ···················· | ···················· | ···················· | ···················· | ···················· | ···················· |
| R | 7347 | hCoV-19/USA/GA-CDC-6466/2020\|EPI_ISL_648002\|2020-0 | ···················· | ···················· | ···················· | ···················· | ···················· | ···················· |
| R | 7348 | hCoV-19/USA/GA-CDC-6467/2020\|EPI_ISL_648003\|2020-0 | ···················· | ···················· | ···················· | ···················· | ···················· | ···················· |
| R | 7349 | hCoV-19/USA/MS-CDC-6516/2020\|EPI_ISL_648004\|2020-0 | ···················· | ···················· | ···················· | ···················· | ···················· | ···················· |
| R | 7350 | hCoV-19/USA/MS-CDC-6546/2020\|EPI_ISL_648005\|2020-0 | ···················· | ···················· | ···················· | ···················· | ···················· | ···················· |
| R | 7351 | hCoV-19/USA/MS-CDC-6549/2020\|EPI_ISL_648006\|2020-0 | ···················· | ···················· | ···················· | ···················· | ···················· | ···················· |
| R | 7352 | hCoV-19/USA/MS-CDC-6552/2020\|EPI_ISL_648007\|2020-0 | ···················· | ···················· | ···················· | ···················· | ···················· | ···················· |
| R | 7353 | hCoV-19/USA/MS-CDC-6553/2020\|EPI_ISL_648008\|2020-0 | ···················· | ···················· | ···················· | ···················· | ···················· | ···················· |
| R | 7354 | hCoV-19/USA/MS-CDC-6554/2020\|EPI_ISL_648009\|2020-0 | ···················· | ···················· | ···················· | ···················· | ···················· | ···················· |
| R | 7355 | hCoV-19/USA/MS-CDC-6557/2020\|EPI_ISL_648010\|2020-0 | ···················· | ···················· | ···················· | ···················· | ···················· | ···················· |
| R | 7356 | hCoV-19/USA/MS-CDC-6558/2020\|EPI_ISL_648011\|2020-0 | ···················· | ···················· | ···················· | ···················· | ···················· | ···················· |
| R | 7357 | hCoV-19/USA/MS-CDC-6560/2020\|EPI_ISL_648012\|2020-0 | ···················· | ···················· | ···················· | ···················· | ···················· | ···················· |
| R | 7358 | hCoV-19/USA/MS-CDC-6561/2020\|EPI_ISL_648013\|2020-0 | ···················· | ···················· | ···················· | ···················· | ···················· | ···················· |
| R | 7359 | hCoV-19/USA/MS-CDC-6562/2020\|EPI_ISL_648014\|2020-0 | ···················· | ···················· | ···················· | ···················· | ···················· | ···················· |
| R | 7360 | hCoV-19/USA/MS-CDC-6563/2020\|EPI_ISL_648015\|2020-0 | ···················· | ···················· | ···················· | ···················· | ···················· | ···················· |
| R | 7361 | hCoV-19/USA/MS-CDC-6564/2020\|EPI_ISL_648016\|2020-0 | ···················· | ···················· | ···················· | ···················· | ···················· | ···················· |
| R | 7362 | hCoV-19/USA/MS-CDC-6565/2020\|EPI_ISL_648017\|2020-0 | ···················· | ···················· | ···················· | ···················· | ···················· | ···················· |
| R | 7363 | hCoV-19/USA/MS-CDC-6566/2020\|EPI_ISL_648018\|2020-0 | ···················· | ···················· | ···················· | ···················· | ···················· | ···················· |
| R | 7364 | hCoV-19/USA/MS-CDC-6567/2020\|EPI_ISL_648019\|2020-0 | ···················· | ···················· | ···················· | ···················· | ···················· | ···················· |
| R | 7365 | hCoV-19/USA/MS-CDC-6568/2020\|EPI_ISL_648020\|2020-0 | ···················· | ···················· | ···················· | ···················· | ···················· | ···················· |
| R | 7366 | hCoV-19/USA/MS-CDC-6569/2020\|EPI_ISL_648021\|2020-0 | ···················· | ···················· | ···················· | ···················· | ···················· | ···················· |
| R | 7367 | hCoV-19/USA/MS-CDC-6570/2020\|EPI_ISL_648022\|2020-0 | ···················· | ···················· | ···················· | ···················· | ···················· | ···················· |
| R | 7368 | hCoV-19/USA/MS-CDC-6571/2020\|EPI_ISL_648023\|2020-0 | ···················· | ···················· | ···················· | ···················· | ···················· | ···················· |
| R | 7369 | hCoV-19/USA/MS-CDC-6572/2020\|EPI_ISL_648024\|2020-0 | ···················· | ···················· | ···················· | ···················· | ···················· | ···················· |
| R | 7370 | hCoV-19/USA/MS-CDC-6573/2020\|EPI_ISL_648025\|2020-0 | ···················· | ···················· | ···················· | ···················· | ···················· | ···················· |
| R | 7371 | hCoV-19/USA/MS-CDC-6574/2020\|EPI_ISL_648026\|2020-0 | ···················· | ···················· | ···················· | ···················· | ···················· | ···················· |
| R | 7372 | hCoV-19/USA/MS-CDC-6575/2020\|EPI_ISL_648027\|2020-0 | ···················· | ···················· | ···················· | ···················· | ···················· | ···················· |
| R | 7373 | hCoV-19/USA/MS-CDC-6576/2020\|EPI_ISL_648028\|2020-0 | ···················· | ···················· | ···················· | ···················· | ···················· | ···················· |
| R | 7374 | hCoV-19/USA/MS-CDC-6577/2020\|EPI_ISL_648029\|2020-0 | ···················· | ···················· | ···················· | ···················· | ···················· | ···················· |
| R | 7375 | hCoV-19/USA/MS-CDC-6579/2020\|EPI_ISL_648030\|2020-0 | ···················· | ···················· | ···················· | ···················· | ···················· | ···················· |
| R | 7376 | hCoV-19/USA/MS-CDC-6580/2020\|EPI_ISL_648031\|2020-0 | ···················· | ···················· | ···················· | ···················· | ···················· | ···················· |
| R | 7377 | hCoV-19/USA/MS-CDC-6581/2020\|EPI_ISL_648032\|2020-0 | ···················· | ···················· | ···················· | ···················· | ···················· | ···················· |
| R | 7378 | hCoV-19/USA/MS-CDC-6583/2020\|EPI_ISL_648033\|2020-0 | ···················· | ···················· | ···················· | ···················· | ···················· | ···················· |
| R | 7379 | hCoV-19/USA/MS-CDC-6585/2020\|EPI_ISL_648034\|2020-0 | ···················· | ···················· | ···················· | ···················· | ···················· | ···················· |
| R | 7380 | hCoV-19/USA/WI-CDC-8095/2020\|EPI_ISL_648035\|2020-0 | ···················· | ···················· | ···················· | ···················· | ···················· | ···················· |
| R | 7381 | hCoV-19/USA/WI-CDC-8097/2020\|EPI_ISL_648036\|2020-0 | ···················· | ···················· | ···················· | ···················· | ···················· | ···················· |
| R | 7382 | hCoV-19/USA/MS-CDC-6513/2020\|EPI_ISL_648037\|2020-0 | ···················· | ·A·················· | ···················· | ···················· | ···················· | ···················· |
| R | 7383 | hCoV-19/USA/MS-CDC-6515/2020\|EPI_ISL_648038\|2020-0 | ···················· | ···················· | ···················· | ···················· | ···················· | ···················· |
| R | 7384 | hCoV-19/USA/MS-CDC-6517/2020\|EPI_ISL_648039\|2020-0 | ···················· | ···················· | ···················· | ···················· | ···················· | ···················· |
| R | 7385 | hCoV-19/USA/MS-CDC-6556/2020\|EPI_ISL_648040\|2020-0 | ···················· | ···················· | ···················· | ···················· | ···················· | ···················· |
| R | 7386 | hCoV-19/USA/MS-CDC-6584/2020\|EPI_ISL_648041\|2020-0 | ···················· | ···················· | ···················· | ···················· | ···················· | ···················· |
| R | 7387 | hCoV-19/USA/MS-CDC-6578/2020\|EPI_ISL_648042\|2020-0 | ···················· | ···················· | ···················· | ···················· | ···················· | ···················· |
| R | 7388 | hCoV-19/Singapore/1135/2020\|EPI_ISL_648043\|2020-02 | ···················· | ···················· | ···················· | ···················· | ···················· | ···················· |
| R | 7389 | hCoV-19/Singapore/37nan/2020\|EPI_ISL_648044\|2020-0 | ···················· | ···················· | ···················· | ···················· | ···················· | ···················· |
| R | 7390 | hCoV-19/Singapore/1136/2020\|EPI_ISL_648045\|2020-03 | ···················· | ···················· | ···················· | ···················· | ···················· | ···················· |
| R | 7391 | hCoV-19/Singapore/1137/2020\|EPI_ISL_648046\|2020-04 | ···················· | ···················· | ···················· | ···················· | ···················· | ···················· |
| R | 7392 | hCoV-19/Singapore/1138/2020\|EPI_ISL_648047\|2020-04 | ···················· | ···················· | ···················· | ···················· | ···················· | ···················· |
| R | 7393 | hCoV-19/Singapore/1139/2020\|EPI_ISL_648048\|2020-04 | ···················· | ···················· | ···················· | ···················· | ···················· | ···················· |
| R | 7394 | hCoV-19/Singapore/1140/2020\|EPI_ISL_648049\|2020-04 | ···················· | ···················· | ···················· | ···················· | ···················· | ···················· |
| R | 7395 | hCoV-19/Singapore/1141/2020\|EPI_ISL_648050\|2020-04 | ···················· | ···················· | ···················· | ···················· | ···················· | ···················· |
| R | 7396 | hCoV-19/Singapore/1142/2020\|EPI_ISL_648051\|2020-04 | ···················· | ···················· | ···················· | ···················· | ···················· | ···················· |
| R | 7397 | hCoV-19/Singapore/1143/2020\|EPI_ISL_648052\|2020-04 | ···················· | ···················· | ···················· | ···················· | ···················· | ···················· |
| R | 7398 | hCoV-19/Singapore/1144/2020\|EPI_ISL_648053\|2020-04 | ···················· | ···················· | ···················· | ···················· | ···················· | ···················· |
| R | 7399 | hCoV-19/Singapore/1145/2020\|EPI_ISL_648054\|2020-04 | ···················· | ···················· | ···················· | ···················· | ···················· | ···················· |
| R | 7400 | hCoV-19/Singapore/1146/2020\|EPI_ISL_648055\|2020-04 | ···················· | ···················· | ···················· | ···················· | ···················· | ···················· |
| R | 7401 | hCoV-19/Singapore/1147/2020\|EPI_ISL_648056\|2020-04 | ···················· | ···················· | ···················· | ···················· | ···················· | ···················· |
| R | 7402 | hCoV-19/Singapore/1148/2020\|EPI_ISL_648057\|2020-04 | ···················· | ···················· | ···················· | ···················· | ···················· | ···················· |
| R | 7403 | hCoV-19/Singapore/1149/2020\|EPI_ISL_648058\|2020-04 | ···················· | ···················· | ···················· | ···················· | ···················· | ···················· |
| R | 7404 | hCoV-19/Singapore/1150/2020\|EPI_ISL_648059\|2020-04 | ···················· | ···················· | ···················· | ···················· | ···················· | ···················· |
| R | 7405 | hCoV-19/Singapore/1151/2020\|EPI_ISL_648060\|2020-04 | ···················· | ···················· | ···················· | ···················· | ···················· | ···················· |
| R | 7406 | hCoV-19/Singapore/1152/2020\|EPI_ISL_648061\|2020-04 | ···················· | ···················· | ···················· | ···················· | ···················· | ···················· |
| R | 7407 | hCoV-19/Singapore/34nan/2020\|EPI_ISL_648062\|2020-0 | ···················· | ···················· | ···················· | ···················· | ···················· | ···················· |
| R | 7408 | hCoV-19/Singapore/106nan/2020\|EPI_ISL_648063\|2020- | ···················· | ···················· | ···················· | ···················· | ···················· | ···················· |
| R | 7409 | hCoV-19/Singapore/139nan/2020\|EPI_ISL_648064\|2020- | ···················· | ···················· | ···················· | ···················· | ···················· | ···················· |
| R | 7410 | hCoV-19/Singapore/433nan/2020\|EPI_ISL_648065\|2020- | ···················· | ···················· | ···················· | ···················· | ···················· | ···················· |
| R | 7411 | hCoV-19/Singapore/141nan/2020\|EPI_ISL_648066\|2020- | ···················· | ···················· | ···················· | ···················· | ···················· | ···················· |
| R | 7412 | hCoV-19/Singapore/1153/2020\|EPI_ISL_648067\|2020-03 | ···················· | ···················· | ···················· | ···················· | ···················· | ···················· |
| R | 7413 | hCoV-19/Singapore/985nan/2020\|EPI_ISL_648068\|2020- | ···················· | ···················· | ···················· | ···················· | ···················· | ···················· |
| R | 7414 | hCoV-19/Singapore/132nan/2020\|EPI_ISL_648069\|2020- | ···················· | ···················· | ···················· | ···················· | ···················· | ···················· |
| R | 7415 | hCoV-19/Singapore/167nan/2020\|EPI_ISL_648070\|2020- | ···················· | ···················· | ···················· | ···················· | ···················· | ···················· |
| R | 7416 | hCoV-19/Singapore/17nan/2020\|EPI_ISL_648071\|2020-0 | ···················· | ···················· | ···················· | ···················· | ···················· | ···················· |
| R | 7417 | hCoV-19/Singapore/1154/2020\|EPI_ISL_648072\|2020-03 | ···················· | ···················· | ···················· | ···················· | ···················· | ···················· |
| R | 7418 | hCoV-19/Singapore/435nan/2020\|EPI_ISL_648073\|2020- | ···················· | ···················· | ···················· | ···················· | ···················· | ···················· |

| | | | | | | | | |
|---|---|---|---|---|---|---|---|---|
| R | 7419 | hCoV-19/Singapore/1406/2020\|EPI_ISL_648074\|2020-03 | ···················· | ···················· | ···················· | ···················· | ···················· | ···················· |
| R | 7420 | hCoV-19/Singapore/146nan/2020\|EPI_ISL_648075\|2020- | ···················· | ···················· | ···················· | ···················· | ···················· | ···················· |
| R | 7421 | hCoV-19/Singapore/1407/2020\|EPI_ISL_648076\|2020-03 | ···················· | ···················· | ···················· | ···················· | ···················· | ···················· |
| R | 7422 | hCoV-19/Singapore/110nan/2020\|EPI_ISL_648077\|2020- | ···················· | ···················· | ···················· | ···················· | ···················· | ···················· |
| R | 7423 | hCoV-19/Singapore/287nan/2020\|EPI_ISL_648078\|2020- | ···················· | ···················· | ···················· | ···················· | ···················· | ···················· |
| R | 7424 | hCoV-19/Singapore/94nan/2020\|EPI_ISL_648079\|2020-0 | ···················· | ···················· | ···················· | ···················· | ···················· | ···················· |
| R | 7425 | hCoV-19/Ireland/MN-NVRL-20IRL51408/2020\|EPI_ISL_63 | ···················· | ···················· | ···················· | ···················· | ···················· | ···················· |
| R | 7426 | hCoV-19/Ireland/CN-NVRL-20IRL51356/2020\|EPI_ISL_63 | ···················· | ···················· | ···················· | ···················· | ···················· | ···················· |
| R | 7427 | hCoV-19/Ireland/MN-NVRL-20IRL52459/2020\|EPI_ISL_63 | ···················· | ···················· | ···················· | ···················· | ···················· | ···················· |
| R | 7428 | hCoV-19/Russia/OMS-ORINFI-1123S/2020\|EPI_ISL_63991 | ···················· | ···················· | ···················· | ···················· | ···················· | ···················· |
| R | 7429 | hCoV-19/Russia/OMS-ORINFI-1564S/2020\|EPI_ISL_63991 | ···················· | ···················· | ···················· | ···················· | ···················· | ···················· |
| R | 7430 | hCoV-19/Russia/OMS-ORINFI-1612S/2020\|EPI_ISL_63991 | ···················· | ···················· | ···················· | ···················· | ···················· | ···················· |
| R | 7431 | hCoV-19/Russia/OMS-ORINFI-1068S/2020\|EPI_ISL_63991 | ···················· | ···················· | ···················· | ···················· | ···················· | ···················· |
| R | 7432 | hCoV-19/Russia/OMS-ORINFI-2087S/2020\|EPI_ISL_63991 | ···················· | ···················· | ···················· | ···················· | ···················· | ···················· |
| R | 7433 | hCoV-19/Russia/OMS-ORINFI-2096S/2020\|EPI_ISL_63991 | ···················· | ···················· | ···················· | ···················· | ···················· | ···················· |
| R | 7434 | hCoV-19/Russia/OMS-ORINFI-2114S/2020\|EPI_ISL_63991 | ···················· | ···················· | ···················· | ···················· | ···················· | ···················· |
| R | 7435 | hCoV-19/Russia/OMS-ORINFI-2166S/2020\|EPI_ISL_63992 | ···················· | ···················· | ···················· | ···················· | ···················· | ···················· |
| R | 7436 | hCoV-19/Russia/OMS-ORINFI-2292S/2020\|EPI_ISL_63992 | ···················· | ···················· | ···················· | ···················· | ···················· | ···················· |
| R | 7437 | hCoV-19/Russia/OMS-ORINFI-2307S/2020\|EPI_ISL_63992 | ···················· | ···················· | ···················· | ···················· | ···················· | ···················· |
| R | 7438 | hCoV-19/Russia/OMS-ORINFI-3769S/2020\|EPI_ISL_63992 | ···················· | ···················· | ···················· | ···················· | ···················· | ···················· |
| R | 7439 | hCoV-19/Russia/OMS-ORINFI-3801S/2020\|EPI_ISL_63992 | ···················· | ···················· | ···················· | ···················· | ···················· | ···················· |
| R | 7440 | hCoV-19/Russia/OMS-ORINFI-3816S/2020\|EPI_ISL_63992 | ···················· | ···················· | ···················· | ···················· | ···················· | ···················· |
| R | 7441 | hCoV-19/Russia/OMS-ORINFI-3818S/2020\|EPI_ISL_63992 | ···················· | ···················· | ···················· | ···················· | ···················· | ···················· |
| R | 7442 | hCoV-19/Australia/VIC12792/2020\|EPI_ISL_640901\|202 | ···················· | ···················· | ···················· | ···················· | ···················· | ···················· |
| R | 7443 | hCoV-19/Australia/VIC16096/2020\|EPI_ISL_640903\|202 | ···················· | ···················· | ···················· | ···················· | ···················· | ···················· |
| R | 7444 | hCoV-19/Australia/VIC12683/2020\|EPI_ISL_640904\|202 | ···················· | ·····A·············· | ···················· | ···················· | ···················· | ···················· |
| R | 7445 | hCoV-19/Australia/VIC12354/2020\|EPI_ISL_640905\|202 | ···················· | ···················· | ···················· | ···················· | ···················· | ···················· |
| R | 7446 | hCoV-19/Australia/VIC16007/2020\|EPI_ISL_640906\|202 | ···················· | ···················· | ···················· | ···················· | ···················· | ···················· |
| R | 7447 | hCoV-19/Australia/VIC16392/2020\|EPI_ISL_640907\|202 | ···················· | ···················· | ···················· | ···················· | ···················· | ···················· |
| R | 7448 | hCoV-19/Australia/VIC12002/2020\|EPI_ISL_640908\|202 | ···················· | ···················· | ···················· | ···················· | ···················· | ···················· |
| R | 7449 | hCoV-19/Australia/VIC11805/2020\|EPI_ISL_640909\|202 | ···················· | ···················· | ···················· | ···················· | ···················· | ···················· |
| R | 7450 | hCoV-19/England/QEUH-A8B66B/2020\|EPI_ISL_643106\|20 | ···················· | ···················· | ···················· | ···················· | ···················· | ···················· |
| R | 7451 | hCoV-19/England/QEUH-A8CA13/2020\|EPI_ISL_643107\|20 | ···················· | ···················· | ···················· | ···················· | ···················· | ···················· |
| R | 7452 | hCoV-19/England/QEUH-A8C7D0/2020\|EPI_ISL_643108\|20 | ···················· | ···················· | ···················· | ···················· | ···················· | ···················· |
| R | 7453 | hCoV-19/England/QEUH-A8C9E9/2020\|EPI_ISL_643109\|20 | ···················· | ···················· | ···················· | ···················· | ···················· | ···················· |
| R | 7454 | hCoV-19/England/QEUH-A8C239/2020\|EPI_ISL_643110\|20 | ···················· | ···················· | ···················· | ···················· | ···················· | ···················· |
| R | 7455 | hCoV-19/England/QEUH-A8C4CA/2020\|EPI_ISL_643111\|20 | ···················· | ···················· | ···················· | ···················· | ···················· | ···················· |
| R | 7456 | hCoV-19/England/QEUH-A8C415/2020\|EPI_ISL_643112\|20 | ···················· | ···················· | ···················· | ···················· | ···················· | ···················· |
| R | 7457 | hCoV-19/England/QEUH-A8C3DC/2020\|EPI_ISL_643114\|20 | ···················· | ···················· | ···················· | ···················· | ···················· | ···················· |
| R | 7458 | hCoV-19/England/QEUH-A8C62E/2020\|EPI_ISL_643115\|20 | ···················· | ···················· | ···················· | ···················· | ···················· | ···················· |
| R | 7459 | hCoV-19/England/QEUH-A8C381/2020\|EPI_ISL_643116\|20 | ···················· | ···················· | ···················· | ···················· | ···················· | ···················· |
| R | 7460 | hCoV-19/England/QEUH-A8BB5D/2020\|EPI_ISL_643117\|20 | ···················· | ···················· | ···················· | ···················· | ···················· | ···················· |
| R | 7461 | hCoV-19/England/QEUH-A8B346/2020\|EPI_ISL_643118\|20 | ···················· | ···················· | ···················· | ···················· | ···················· | ···················· |
| R | 7462 | hCoV-19/England/QEUH-A8B856/2020\|EPI_ISL_643119\|20 | ···················· | ···················· | ···················· | ···················· | ···················· | ···················· |
| R | 7463 | hCoV-19/England/QEUH-A8C65B/2020\|EPI_ISL_643121\|20 | ···················· | ···················· | ···················· | ···················· | ···················· | ···················· |
| R | 7464 | hCoV-19/England/QEUH-A8C327/2020\|EPI_ISL_643122\|20 | ···················· | ···················· | ···················· | ···················· | ···················· | ···················· |
| R | 7465 | hCoV-19/England/QEUH-A8BFCA/2020\|EPI_ISL_643123\|20 | ···················· | ···················· | ···················· | ···················· | ···················· | ···················· |
| R | 7466 | hCoV-19/England/QEUH-A8BC69/2020\|EPI_ISL_643124\|20 | ···················· | ···················· | ···················· | ···················· | ···················· | ···················· |
| R | 7467 | hCoV-19/England/QEUH-A8B30A/2020\|EPI_ISL_643125\|20 | ···················· | ···················· | ···················· | ···················· | ···················· | ···················· |
| R | 7468 | hCoV-19/England/QEUH-A8BD0C/2020\|EPI_ISL_643126\|20 | ···················· | ···················· | ···················· | ···················· | ···················· | ···················· |
| R | 7469 | hCoV-19/England/QEUH-A8BF60/2020\|EPI_ISL_643127\|20 | ···················· | ···················· | ···················· | ················G··· | ···················· | ···················· |
| R | 7470 | hCoV-19/England/QEUH-A8C460/2020\|EPI_ISL_643129\|20 | ···················· | ···················· | ···················· | ···················· | ···················· | ···················· |
| R | 7471 | hCoV-19/England/QEUH-A8BBC6/2020\|EPI_ISL_643130\|20 | ···················· | ···················· | ···················· | ···················· | ···················· | ···················· |
| R | 7472 | hCoV-19/England/QEUH-A8BE81/2020\|EPI_ISL_643131\|20 | ···················· | ···················· | ···················· | ···················· | ···················· | ···················· |
| R | 7473 | hCoV-19/England/QEUH-A8C47F/2020\|EPI_ISL_643132\|20 | ···················· | ···················· | ···················· | ···················· | ···················· | ···················· |
| R | 7474 | hCoV-19/England/QEUH-A8BFF7/2020\|EPI_ISL_643133\|20 | ···················· | ···················· | ···················· | ···················· | ···················· | ···················· |
| R | 7475 | hCoV-19/England/QEUH-A8B382/2020\|EPI_ISL_643134\|20 | ···················· | ···················· | ···················· | ···················· | ···················· | ···················· |
| R | 7476 | hCoV-19/England/QEUH-A8C178/2020\|EPI_ISL_643135\|20 | ···················· | ···················· | ···················· | ···················· | ···················· | ···················· |
| R | 7477 | hCoV-19/England/QEUH-A8C8DD/2020\|EPI_ISL_643136\|20 | ···················· | ···················· | ···················· | ···················· | ···················· | ···················· |
| R | 7478 | hCoV-19/England/QEUH-A8C61F/2020\|EPI_ISL_643137\|20 | ···················· | ···················· | ···················· | ···················· | ···················· | ···················· |
| R | 7479 | hCoV-19/England/QEUH-A8C697/2020\|EPI_ISL_643139\|20 | ···················· | ···················· | ···················· | ···················· | ···················· | ···················· |
| R | 7480 | hCoV-19/England/QEUH-A8C882/2020\|EPI_ISL_643140\|20 | ···················· | ···················· | ···················· | ···················· | ···················· | ···················· |
| R | 7481 | hCoV-19/England/QEUH-A8B48F/2020\|EPI_ISL_643142\|20 | ···················· | ···················· | ···················· | ···················· | ···················· | ···················· |
| R | 7482 | hCoV-19/England/QEUH-A8BA32/2020\|EPI_ISL_643143\|20 | ···················· | ···················· | ···················· | ···················· | ···················· | ···················· |
| R | 7483 | hCoV-19/England/QEUH-A8C293/2020\|EPI_ISL_643144\|20 | ···················· | ···················· | ···················· | ···················· | ···················· | ···················· |
| R | 7484 | hCoV-19/England/QEUH-A8B71D/2020\|EPI_ISL_643145\|20 | ···················· | ···················· | ···················· | ···················· | ···················· | ···················· |
| R | 7485 | hCoV-19/England/QEUH-A8B7B3/2020\|EPI_ISL_643146\|20 | ···················· | ···················· | ···················· | ···················· | ···················· | ···················· |
| R | 7486 | hCoV-19/England/QEUH-A8C819/2020\|EPI_ISL_643147\|20 | ···················· | ···················· | ···················· | ···················· | ···················· | ···················· |
| R | 7487 | hCoV-19/England/QEUH-A8BA23/2020\|EPI_ISL_643148\|20 | ···················· | ···················· | ···················· | ···················· | ···················· | ···················· |
| R | 7488 | hCoV-19/England/QEUH-A8C4D9/2020\|EPI_ISL_643150\|20 | ···················· | ···················· | ···················· | ···················· | ···················· | ···················· |
| R | 7489 | hCoV-19/England/QEUH-A8C679/2020\|EPI_ISL_643151\|20 | ···················· | ···················· | ···················· | ···················· | ···················· | ···················· |
| R | 7490 | hCoV-19/England/QEUH-A8B285/2020\|EPI_ISL_643152\|20 | ···················· | ···················· | ···················· | ···················· | ···················· | ···················· |
| R | 7491 | hCoV-19/England/QEUH-A8B953/2020\|EPI_ISL_643153\|20 | ···················· | ···················· | ···················· | ···················· | ···················· | ···················· |
| R | 7492 | hCoV-19/England/QEUH-A8C3AF/2020\|EPI_ISL_643154\|20 | ···················· | ···················· | ···················· | ···················· | ···················· | ···················· |
| R | 7493 | hCoV-19/England/QEUH-A8B57D/2020\|EPI_ISL_643155\|20 | ···················· | ···················· | ···················· | ···················· | ···················· | ···················· |
| R | 7494 | hCoV-19/England/QEUH-A8C3BE/2020\|EPI_ISL_643156\|20 | ···················· | ···················· | ···················· | ···················· | ···················· | ···················· |
| R | 7495 | hCoV-19/England/QEUH-A8C916/2020\|EPI_ISL_643157\|20 | ···················· | ···················· | ···················· | ···················· | ···················· | ···················· |
| R | 7496 | hCoV-19/England/QEUH-A8C1E1/2020\|EPI_ISL_643158\|20 | ···················· | ···················· | ···················· | ···················· | ···················· | ···················· |
| R | 7497 | hCoV-19/England/QEUH-A8C8BF/2020\|EPI_ISL_643159\|20 | ···················· | ···················· | ···················· | ···················· | ···················· | ···················· |
| R | 7498 | hCoV-19/England/QEUH-A8BBA8/2020\|EPI_ISL_643160\|20 | ···················· | ···················· | ···················· | ···················· | ···················· | ···················· |

R 7499 hCoV-19/England/QEUH-A8B4BC/2020|EPI_ISL_643161|20 ···················· ···················· ···················· ···················· ···················· ····················
R 7500 hCoV-19/England/QEUH-A8BCE1/2020|EPI_ISL_643162|20 ···················· ···················· ···················· ···················· ···················· ····················
R 7501 hCoV-19/England/QEUH-A8BA05/2020|EPI_ISL_643163|20 ···················· ···················· ···················· ···················· ···················· ····················
R 7502 hCoV-19/England/QEUH-A8B470/2020|EPI_ISL_643164|20 ···················· ···················· ···················· ···················· ···················· ····················
R 7503 hCoV-19/England/QEUH-A8C7EF/2020|EPI_ISL_643165|20 ···················· ···················· ···················· ···················· ···················· ····················
R 7504 hCoV-19/England/QEUH-A8BCB4/2020|EPI_ISL_643166|20 ···················· ···················· ···················· ···················· ···················· ····················
R 7505 hCoV-19/England/QEUH-A8B461/2020|EPI_ISL_643167|20 ···················· ···················· ···················· ···················· ···················· ····················
R 7506 hCoV-19/England/QEUH-A8B74A/2020|EPI_ISL_643168|20 ···················· ···················· ···················· ···················· ···················· ····················
R 7507 hCoV-19/England/QEUH-A8C828/2020|EPI_ISL_643169|20 ···················· ···················· ···················· ···················· ···················· ····················
R 7508 hCoV-19/England/QEUH-A8B531/2020|EPI_ISL_643176|20 ···················· ···················· ···················· ···················· ···················· ····················
R 7509 hCoV-19/England/QEUH-A8B416/2020|EPI_ISL_643177|20 ···················· ···················· ···················· ···················· ···················· ····················
R 7510 hCoV-19/England/QEUH-A8B926/2020|EPI_ISL_643178|20 ···················· ···················· ···················· ···················· ···················· ····················
R 7511 hCoV-19/England/QEUH-A8C4F7/2020|EPI_ISL_643181|20 ···················· ···················· ···················· ···················· ···················· ····················
R 7512 hCoV-19/England/QEUH-A8C512/2020|EPI_ISL_643182|20 ···················· ···················· ···················· ···················· ···················· ····················
R 7513 hCoV-19/England/QEUH-A8C451/2020|EPI_ISL_643183|20 ···················· ···················· ···················· ···················· ···················· ····················
R 7514 hCoV-19/England/QEUH-A8C4AC/2020|EPI_ISL_643184|20 ···················· ···················· ···················· ···················· ···················· ····················
R 7515 hCoV-19/England/QEUH-A8BC96/2020|EPI_ISL_643185|20 ···················· ···················· ···················· ···················· ···················· ····················
R 7516 hCoV-19/England/QEUH-A8B980/2020|EPI_ISL_643186|20 ···················· ···················· ···················· ···················· ···················· ····················
R 7517 hCoV-19/England/QEUH-A8B9BD/2020|EPI_ISL_643187|20 ···················· ·A·················· ···················· ···················· ···················· ···A················
R 7518 hCoV-19/England/QEUH-A8BC3C/2020|EPI_ISL_643188|20 ···················· ···················· ···················· ···················· ···················· ····················
R 7519 hCoV-19/England/QEUH-A8C372/2020|EPI_ISL_643189|20 ···················· ···················· ···················· ···················· ···················· ····················
R 7520 hCoV-19/France/PAC-MEPHI-1736/2020|EPI_ISL_644437| ···················· ···················· ···················· ···················· ···················· ····················
R 7521 hCoV-19/France/PAC-MEPHI-1738/2020|EPI_ISL_644438| ···················· ···················· ···················· ···················· ···················· ····················
R 7522 hCoV-19/France/PAC-MEPHI-1741/2020|EPI_ISL_644439| ···················· ···················· ···················· ···················· ···················· ····················
R 7523 hCoV-19/France/PAC-MEPHI-1746/2020|EPI_ISL_644440| ···················· ···················· ···················· ···················· ···················· ····················
R 7524 hCoV-19/France/PAC-MEPHI-1749/2020|EPI_ISL_644441| ···················· ···················· ···················· ···················· ···················· ····················
R 7525 hCoV-19/France/PAC-MEPHI-1755/2020|EPI_ISL_644442| ···················· ···················· ···················· ···················· ···················· ····················
R 7526 hCoV-19/France/PAC-MEPHI-1758/2020|EPI_ISL_644443| ···················· ···················· ···················· ···················· ···················· ····················
R 7527 hCoV-19/France/PAC-MEPHI-1762/2020|EPI_ISL_644444| ···················· ···················· ···················· ···················· ···················· ····················
R 7528 hCoV-19/France/PAC-MEPHI-1764/2020|EPI_ISL_644445| ···················· ···················· ···················· ···················· ···················· ····················
R 7529 hCoV-19/France/PAC-MEPHI-1771/2020|EPI_ISL_644446| ···················· ···················· ···················· ···················· ···················· ····················
R 7531 hCoV-19/France/PAC-MEPHI-1882/2020|EPI_ISL_644448| ···················· ···················· ···················· ···················· ···················· ····················
R 7532 hCoV-19/France/PAC-MEPHI-1883/2020|EPI_ISL_644449| ···················· ···················· ···················· ···················· ···················· ····················
R 7533 hCoV-19/France/PAC-MEPHI-1884/2020|EPI_ISL_644450| ···················· ···················· ···················· ···················· ···················· ····················
R 7534 hCoV-19/France/PAC-MEPHI-1886/2020|EPI_ISL_644451| ···················· ···················· ···················· ···················· ···················· ····················
R 7535 hCoV-19/France/PAC-MEPHI-1897/2020|EPI_ISL_644452| ···················· ···················· ···················· ···················· ···················· ····················
R 7536 hCoV-19/France/PAC-MEPHI-1899/2020|EPI_ISL_644453| ···················· ···················· ···················· ···················· ···················· ····················
R 7537 hCoV-19/France/PAC-MEPHI-1902/2020|EPI_ISL_644454| ···················· ···A················ ···················· ···················· ···················· ····················
R 7538 hCoV-19/France/PAC-MEPHI-1905/2020|EPI_ISL_644455| ···················· ···················· ···················· ···················· ···················· ····················
R 7539 hCoV-19/France/PAC-MEPHI-1907/2020|EPI_ISL_644456| ···················· ···················· ···················· ···················· ···················· ····················
R 7540 hCoV-19/France/PAC-MEPHI-1908/2020|EPI_ISL_644457| ···················· ···················· ···················· ···················· ···················· ····················
R 7541 hCoV-19/France/PAC-MEPHI-1911/2020|EPI_ISL_644458| ···················· ···················· ···················· ···················· ···················· ····················
R 7542 hCoV-19/France/PAC-MEPHI-1912/2020|EPI_ISL_644459| ···················· ···················· ···················· ···················· ···················· ····················
R 7543 hCoV-19/France/PAC-MEPHI-1914/2020|EPI_ISL_644460| ···················· ···················· ···················· ···················· ···················· ····················
R 7544 hCoV-19/France/PAC-MEPHI-1927/2020|EPI_ISL_644461| ···················· ···················· ···················· ···················· ···················· ····················
R 7545 hCoV-19/France/PAC-MEPHI-1929/2020|EPI_ISL_644462| ···················· ·A·················· ···················· ···················· ···················· ····················
R 7546 hCoV-19/France/PAC-MEPHI-1931/2020|EPI_ISL_644463| ···················· ···················· ···················· ···················· ···················· ····················
R 7547 hCoV-19/France/PAC-MEPHI-1935/2020|EPI_ISL_644464| ···················· ···················· ···················· ···················· ···················· ····················
R 7548 hCoV-19/France/PAC-MEPHI-1940/2020|EPI_ISL_644465| ···················· ···················· ···················· ···················· ···················· ····················
R 7549 hCoV-19/France/PAC-MEPHI-1972/2020|EPI_ISL_644466| ···················· ···················· ···················· ···················· ···················· ····················
R 7550 hCoV-19/France/PAC-MEPHI-1976/2020|EPI_ISL_644467| ···················· ···················· ···················· ···················· ···················· ····················
R 7551 hCoV-19/France/PAC-MEPHI-1980/2020|EPI_ISL_644468| ···················· ···················· ···················· ···················· ···················· ····················
R 7552 hCoV-19/France/PAC-MEPHI-1989/2020|EPI_ISL_644469| ···················· ···················· ···················· ···················· ···················· ····················
R 7553 hCoV-19/France/PAC-MEPHI-1999/2020|EPI_ISL_644470| ···················· ···················· ···················· ···················· ···················· ····················
R 7554 hCoV-19/France/PAC-MEPHI-2010/2020|EPI_ISL_644471| ···················· ···················· ···················· ···················· ···················· ····················
R 7555 hCoV-19/France/PAC-MEPHI-2011/2020|EPI_ISL_644472| ···················· ···················· ···················· ···················· ···················· ····················
R 7556 hCoV-19/France/PAC-MEPHI-2012/2020|EPI_ISL_644473| ···················· ···················· ···················· ···················· ···················· ····················
R 7557 hCoV-19/France/PAC-MEPHI-2014/2020|EPI_ISL_644474| ···················· ···················· ···················· ···················· ···················· ····················
R 7558 hCoV-19/France/PAC-MEPHI-2018/2020|EPI_ISL_644475| ···················· ···················· ···················· ···················· ···················· ····················
R 7559 hCoV-19/France/PAC-MEPHI-2019/2020|EPI_ISL_644476| ···················· ···················· ···················· ···················· ···················· ····················
R 7560 hCoV-19/France/PAC-MEPHI-2023/2020|EPI_ISL_644477| ···················· ···················· ···················· ···················· ···················· ····················
R 7561 hCoV-19/France/PAC-MEPHI-2033/2020|EPI_ISL_644478| ···················· ···················· ···················· ···················· ···················· ····················
R 7562 hCoV-19/France/PAC-MEPHI-2038/2020|EPI_ISL_644479| ···················· ···················· ···················· ···················· ···················· ····················
R 7563 hCoV-19/France/PAC-MEPHI-2041/2020|EPI_ISL_644480| ···················· ···················· ···················· ···················· ···················· ····················
R 7564 hCoV-19/France/PAC-MEPHI-2049/2020|EPI_ISL_644481| ···················· ···················· ···················· ···················· ···················· ····················
R 7565 hCoV-19/France/PAC-MEPHI-2050/2020|EPI_ISL_644482| ···················· ···················· ···················· ···················· ···················· ····················
R 7566 hCoV-19/France/PAC-MEPHI-2051/2020|EPI_ISL_644483| ···················· ···················· ···················· ···················· ···················· ····················
R 7567 hCoV-19/France/PAC-MEPHI-2054/2020|EPI_ISL_644484| ···················· ···················· ···················· ···················· ···················· ····················
R 7568 hCoV-19/France/PAC-MEPHI-2056/2020|EPI_ISL_644485| ···················· ···················· ···················· ···················· ···················· ····················
R 7569 hCoV-19/France/PAC-MEPHI-2057/2020|EPI_ISL_644486| ···················· ···················· ···················· ···················· ···················· ····················
R 7570 hCoV-19/France/PAC-MEPHI-2061/2020|EPI_ISL_644487| ···················· ···················· ···················· ···················· ···················· ····················
R 7571 hCoV-19/France/PAC-MEPHI-2064/2020|EPI_ISL_644488| ···················· ···················· ···················· ···················· ···················· ····················
R 7572 hCoV-19/France/PAC-MEPHI-2069/2020|EPI_ISL_644489| ···················· ···················· ···················· ···················· ···················· ····················
R 7573 hCoV-19/France/PAC-MEPHI-2071/2020|EPI_ISL_644490| ···················· ···················· ···················· ···················· ······T············· ····················
R 7574 hCoV-19/France/PAC-MEPHI-2083/2020|EPI_ISL_644491| ···················· ···················· ···················· ···················· ···················· ····················
R 7575 hCoV-19/France/PAC-MEPHI-2086/2020|EPI_ISL_644492| ···················· ···················· ···················· ···················· ···················· ····················
R 7576 hCoV-19/France/PAC-MEPHI-2087/2020|EPI_ISL_644493| ···················· ···················· ···················· ···················· ···················· ····················
R 7577 hCoV-19/France/PAC-MEPHI-2094/2020|EPI_ISL_644494| ···················· ···················· ···················· ···················· ···················· ····················
R 7578 hCoV-19/France/PAC-MEPHI-2099/2020|EPI_ISL_644495| ···················· ···················· ···················· ···················· ···················· ····················
R 7579 hCoV-19/France/PAC-MEPHI-2101/2020|EPI_ISL_644496| ···················· ···················· ···················· ···················· ···················· ····················

| | | | | | | | | |
|---|---|---|---|---|---|---|---|---|
| R | 7580 | hCoV-19/France/PAC-MEPHI-2103/2020\|EPI_ISL_644497\| | ···················· | ···················· | ···················· | ···················· | ···················· | ···················· |
| R | 7581 | hCoV-19/France/PAC-MEPHI-2104/2020\|EPI_ISL_644498\| | ···················· | ···················· | ···················· | ···················· | ···················· | ···················· |
| R | 7582 | hCoV-19/France/PAC-MEPHI-2105/2020\|EPI_ISL_644499\| | ···················· | ···················· | ···················· | ···················· | ···················· | ···················· |
| R | 7583 | hCoV-19/England/ALDP-B1A75E/2020\|EPI_ISL_645768\|20 | ···················· | ···················· | ···················· | ···················· | ···················· | ···················· |
| R | 7584 | hCoV-19/England/ALDP-B1C8E0/2020\|EPI_ISL_645769\|20 | ···················· | ···················· | ···················· | ···················· | ···················· | ···················· |
| R | 7585 | hCoV-19/England/ALDP-B1CE93/2020\|EPI_ISL_645770\|20 | ···················· | ···················· | ···················· | ···················· | ···················· | ···················· |
| R | 7586 | hCoV-19/England/ALDP-B1A77C/2020\|EPI_ISL_645771\|20 | ···················· | ···················· | ···················· | ···················· | ······T············· | ···················· |
| R | 7587 | hCoV-19/England/ALDP-B1C394/2020\|EPI_ISL_645772\|20 | ···················· | ···················· | ···················· | ···················· | ···················· | ···················· |
| R | 7588 | hCoV-19/England/ALDP-B1C3EF/2020\|EPI_ISL_645773\|20 | ···················· | ···················· | ···················· | ···················· | ···················· | ···················· |
| R | 7589 | hCoV-19/England/ALDP-B1BBF7/2020\|EPI_ISL_645774\|20 | ···················· | ···················· | ···················· | ···················· | ···················· | ···················· |
| R | 7590 | hCoV-19/England/ALDP-B1B5DB/2020\|EPI_ISL_645775\|20 | ···················· | ···················· | ···················· | ···················· | ···················· | ···················· |
| R | 7591 | hCoV-19/England/ALDP-B1BFDD/2020\|EPI_ISL_645776\|20 | ···················· | ···················· | ···················· | ···················· | ···················· | ···················· |
| R | 7592 | hCoV-19/England/ALDP-B1A69D/2020\|EPI_ISL_645777\|20 | ···················· | ···················· | ···················· | ···················· | ······T············· | ···················· |
| R | 7593 | hCoV-19/England/ALDP-B1C4DD/2020\|EPI_ISL_645778\|20 | ···················· | ···················· | ···················· | ···················· | ···················· | ···················· |
| R | 7594 | hCoV-19/England/ALDP-B1E20E/2020\|EPI_ISL_645779\|20 | ···················· | ···················· | ···················· | ···················· | ···················· | ···················· |
| R | 7595 | hCoV-19/England/ALDP-B1B043/2020\|EPI_ISL_645780\|20 | ···················· | ···················· | ···················· | ···················· | ···················· | ···················· |
| R | 7596 | hCoV-19/England/ALDP-B1BC4F/2020\|EPI_ISL_645781\|20 | ···················· | ···················· | ···················· | ···················· | ···················· | ···················· |
| R | 7597 | hCoV-19/England/ALDP-B1C006/2020\|EPI_ISL_645782\|20 | ···················· | ···················· | ···················· | ···················· | ···················· | ···················· |
| R | 7598 | hCoV-19/England/ALDP-B1B2D4/2020\|EPI_ISL_645783\|20 | ···················· | ···················· | ···················· | ···················· | ···················· | ···················· |
| R | 7599 | hCoV-19/England/ALDP-B1BB51/2020\|EPI_ISL_645784\|20 | ···················· | ···················· | ···················· | ···················· | ···················· | ···················· |
| R | 7600 | hCoV-19/England/ALDP-B1CDF0/2020\|EPI_ISL_645785\|20 | ···················· | ···················· | ···················· | ···················· | ···················· | ···················· |
| R | 7601 | hCoV-19/England/ALDP-B1D0AB/2020\|EPI_ISL_645786\|20 | ···················· | ···················· | ···················· | ···················· | ···················· | ···················· |
| R | 7602 | hCoV-19/England/ALDP-B1BB33/2020\|EPI_ISL_645787\|20 | ···················· | ···················· | ···················· | ···················· | ···················· | ···················· |
| R | 7603 | hCoV-19/England/ALDP-B1C58F/2020\|EPI_ISL_645789\|20 | ···················· | ···················· | ···················· | ···················· | ···················· | ···················· |
| R | 7604 | hCoV-19/England/ALDP-B1AF92/2020\|EPI_ISL_645790\|20 | ···················· | ···················· | ···················· | ···················· | ···················· | ···················· |
| R | 7605 | hCoV-19/England/ALDP-B1B1E6/2020\|EPI_ISL_645791\|20 | ···················· | ···················· | ···················· | ···················· | ···················· | ···················· |
| R | 7606 | hCoV-19/England/ALDP-B1BA72/2020\|EPI_ISL_645792\|20 | ···················· | ···················· | ···················· | ···················· | ···················· | ···················· |
| R | 7607 | hCoV-19/England/ALDP-B1BA54/2020\|EPI_ISL_645793\|20 | ···················· | ···················· | ···················· | ···················· | ···················· | ···················· |
| R | 7608 | hCoV-19/England/ALDP-B1A484/2020\|EPI_ISL_645794\|20 | ···················· | ···················· | ···················· | ···················· | ···················· | ···················· |
| R | 7609 | hCoV-19/England/ALDP-B1CD69/2020\|EPI_ISL_645795\|20 | ···················· | ···················· | ···················· | ···················· | ···················· | ···················· |
| R | 7610 | hCoV-19/England/ALDP-B1A150/2020\|EPI_ISL_645796\|20 | ···················· | ···················· | ···················· | ···················· | ···················· | ···················· |
| R | 7611 | hCoV-19/England/ALDP-B1CE48/2020\|EPI_ISL_645797\|20 | ···················· | ···················· | ···················· | ···················· | ···················· | ···················· |
| R | 7612 | hCoV-19/England/ALDP-B1CCD5/2020\|EPI_ISL_645799\|20 | ···················· | ···················· | ···················· | ···················· | ···················· | ···················· |
| S | 1 | hCoV-19/USA/FL-BPHL-2147/2020\|EPI_ISL_653300\|2020- | ···················· | ···················· | ···················· | ···················· | ···················· | ···················· |
| S | 2 | hCoV-19/USA/FL-BPHL-2149/2020\|EPI_ISL_653302\|2020- | ···················· | ···················· | ···················· | ···················· | ···················· | ···················· |
| S | 3 | hCoV-19/Spain/PV-IBV-98008424/2020\|EPI_ISL_654600\| | ···················· | ···················· | ···················· | ···················· | ···················· | ···················· |
| S | 4 | hCoV-19/Spain/PV-IBV-98008425/2020\|EPI_ISL_654601\| | ···················· | ···················· | ···················· | ···················· | ···················· | ···················· |
| S | 5 | hCoV-19/Spain/PV-IBV-98008428/2020\|EPI_ISL_654602\| | ···················· | ···················· | ···················· | ···················· | ···················· | ···················· |
| S | 6 | hCoV-19/Spain/PV-IBV-98008430/2020\|EPI_ISL_654603\| | ···················· | ···················· | ···················· | ···················· | ···················· | ···················· |
| S | 7 | hCoV-19/Spain/PV-IBV-98008432/2020\|EPI_ISL_654604\| | ···················· | ···················· | ···················· | ···················· | ···················· | ···················· |
| S | 8 | hCoV-19/Spain/PV-IBV-98008435/2020\|EPI_ISL_654605\| | ···················· | ·A·················· | ···················· | ···················· | ···················· | ···················· |
| S | 9 | hCoV-19/Spain/PV-IBV-98008437/2020\|EPI_ISL_654606\| | ···················· | ···················· | ···················· | ···················· | ···················· | ···················· |
| S | 10 | hCoV-19/Spain/PV-IBV-98008439/2020\|EPI_ISL_654607\| | ···················· | ···················· | ···················· | ···················· | ···················· | ···················· |
| S | 11 | hCoV-19/Spain/PV-IBV-98008441/2020\|EPI_ISL_654608\| | ···················· | ···················· | ···················· | ···················· | ···················· | ···················· |
| S | 12 | hCoV-19/Spain/PV-IBV-98008443/2020\|EPI_ISL_654609\| | ···················· | ···················· | ···················· | ···················· | ···················· | ···················· |
| S | 13 | hCoV-19/Spain/PV-IBV-98008447/2020\|EPI_ISL_654610\| | ···················· | ···················· | ···················· | ···················· | ···················· | ···················· |
| S | 14 | hCoV-19/Spain/PV-IBV-98008449/2020\|EPI_ISL_654611\| | ···················· | ···················· | ···················· | ···················· | ···················· | ···················· |
| S | 15 | hCoV-19/Spain/PV-IBV-98008451/2020\|EPI_ISL_654612\| | ···················· | ···················· | ···················· | ···················· | ···················· | ···················· |
| S | 16 | hCoV-19/Spain/PV-IBV-98008453/2020\|EPI_ISL_654613\| | ···················· | ···················· | ···················· | ···················· | ···················· | ···················· |
| S | 17 | hCoV-19/Spain/PV-IBV-98008457/2020\|EPI_ISL_654614\| | ···················· | ·A·················· | ···················· | ···················· | ···················· | ···················· |
| S | 18 | hCoV-19/Spain/PV-IBV-98008461/2020\|EPI_ISL_654615\| | ···················· | ···················· | ···················· | ···················· | ···················· | ···················· |
| S | 19 | hCoV-19/Spain/AS-IBV-98006633/2020\|EPI_ISL_654625\| | ···················· | ···················· | ···················· | ···················· | ···················· | ···················· |
| S | 20 | hCoV-19/Spain/AS-IBV-98006660/2020\|EPI_ISL_654630\| | ···················· | ···················· | ···················· | ···················· | ···················· | ···················· |
| S | 21 | hCoV-19/Spain/AS-IBV-98006628/2020\|EPI_ISL_654631\| | ···················· | ···················· | ···················· | ···················· | ···················· | ···················· |
| S | 22 | hCoV-19/Spain/AS-IBV-98006590/2020\|EPI_ISL_654632\| | ···················· | ···················· | ···················· | ···················· | ···················· | ···················· |
| S | 23 | hCoV-19/Spain/AS-IBV-98006646/2020\|EPI_ISL_654633\| | ···················· | ···················· | ···················· | ···················· | ···················· | ···················· |
| S | 24 | hCoV-19/England/ALDP-B5BE5C/2020\|EPI_ISL_655901\|20 | ···················· | ···················· | ···················· | ···················· | ···················· | ···················· |
| S | 25 | hCoV-19/England/ALDP-B5CC7F/2020\|EPI_ISL_655902\|20 | ···················· | ···················· | ···················· | ···················· | ···················· | ···················· |
| S | 26 | hCoV-19/England/ALDP-B5C644/2020\|EPI_ISL_655903\|20 | ···················· | ···················· | ···················· | ···················· | ···················· | ···················· |
| S | 27 | hCoV-19/England/ALDP-B5C27D/2020\|EPI_ISL_655904\|20 | ···················· | ···················· | ···················· | ···················· | ···················· | ···················· |
| S | 28 | hCoV-19/England/ALDP-B5C7E7/2020\|EPI_ISL_655905\|20 | ···················· | ···················· | ···················· | ···················· | ···················· | ···················· |
| S | 29 | hCoV-19/England/QEUH-B66058/2020\|EPI_ISL_655907\|20 | ···················· | ···················· | ···················· | ···················· | ···················· | ···················· |
| S | 30 | hCoV-19/England/ALDP-B5BEF2/2020\|EPI_ISL_655908\|20 | ···················· | ···················· | ··R················· | ···················· | ···················· | ···················· |
| S | 31 | hCoV-19/England/ALDP-B5C019/2020\|EPI_ISL_655909\|20 | ···················· | ···················· | ···················· | ···················· | ···················· | ···················· |
| S | 32 | hCoV-19/England/ALDP-B5C1DA/2020\|EPI_ISL_655910\|20 | ···················· | ···················· | ···················· | ···················· | ···················· | ···················· |
| S | 33 | hCoV-19/England/QEUH-B6C7B7/2020\|EPI_ISL_655911\|20 | ···················· | ···················· | ···················· | ···················· | ···················· | ···················· |
| S | 34 | hCoV-19/England/QEUH-B6785F/2020\|EPI_ISL_655912\|20 | ···················· | ···················· | ···················· | ···················· | ···················· | ···················· |
| S | 35 | hCoV-19/England/ALDP-B5CBDC/2020\|EPI_ISL_655914\|20 | ···················· | ···················· | ···················· | ···················· | ···················· | ···················· |
| S | 36 | hCoV-19/England/ALDP-B5CCE8/2020\|EPI_ISL_655916\|20 | ···················· | ···················· | ···················· | ···················· | ···················· | ···················· |
| S | 37 | hCoV-19/England/QEUH-B65F10/2020\|EPI_ISL_655918\|20 | ···················· | ·A·················· | ···················· | ···················· | ···················· | ···················· |
| S | 38 | hCoV-19/England/ALDP-B5C7D8/2020\|EPI_ISL_655919\|20 | ···················· | ···················· | ···················· | ···················· | ···················· | ···················· |
| S | 39 | hCoV-19/England/QEUH-B678E6/2020\|EPI_ISL_655920\|20 | ···················· | ···················· | ···················· | ···················· | ···················· | ···················· |
| S | 40 | hCoV-19/England/ALDP-B5C0EC/2020\|EPI_ISL_655921\|20 | ···················· | ·A·················· | ···················· | ···················· | ···················· | ··A················· |
| S | 41 | hCoV-19/England/ALDP-B5C556/2020\|EPI_ISL_655923\|20 | ···················· | ···················· | ···················· | ···················· | ···················· | ···················· |
| S | 42 | hCoV-19/England/QEUH-B67804/2020\|EPI_ISL_655924\|20 | ···················· | ···················· | ···················· | ···················· | ···················· | ···················· |
| S | 43 | hCoV-19/England/ALDP-B5CDA9/2020\|EPI_ISL_655926\|20 | ···················· | ···················· | ···················· | ···················· | ···················· | ···················· |
| S | 44 | hCoV-19/England/ALDP-B5C5FC/2020\|EPI_ISL_655927\|20 | ···················· | ···················· | ···················· | ···················· | ···················· | ···················· |
| S | 45 | hCoV-19/England/ALDP-B5C8B7/2020\|EPI_ISL_655928\|20 | ···················· | ···················· | ···················· | ···················· | ···················· | ···················· |
| S | 46 | hCoV-19/England/QEUH-B6C78A/2020\|EPI_ISL_655929\|20 | ···················· | ···················· | ···················· | ···················· | ···················· | ···················· |
| S | 47 | hCoV-19/England/ALDP-B5C6BD/2020\|EPI_ISL_655930\|20 | ···················· | ···················· | ···················· | ···················· | ···················· | ···················· |

| | | | | | | | |
|---|---|---|---|---|---|---|---|
| S | 48 hCoV-19/England/ALDP-B5C2B9/2020\|EPI_ISL_655931\|20 | ···················· | ···················· | ····················· | ···················· | ···················· | ···················· |
| S | 49 hCoV-19/England/ALDP-B5C477/2020\|EPI_ISL_655932\|20 | ···················· | ···················· | ····················· | ···················· | ···················· | ···················· |
| S | 50 hCoV-19/England/ALDP-B5C33E/2020\|EPI_ISL_655935\|20 | ···················· | ···················· | ····················· | ···················· | ···················· | ···················· |
| S | 51 hCoV-19/England/ALDP-B5C398/2020\|EPI_ISL_655936\|20 | ···················· | ···················· | ····················· | ···················· | ···················· | ···················· |
| S | 52 hCoV-19/England/ALDP-B5CDC7/2020\|EPI_ISL_655937\|20 | ···················· | ···················· | ····················· | ···················· | ···················· | ···················· |
| S | 53 hCoV-19/England/ALDP-B5BCBC/2020\|EPI_ISL_655940\|20 | ···················· | ···················· | ····················· | ···················· | ·····T·············· | ···················· |
| S | 54 hCoV-19/England/ALDP-B5C0BF/2020\|EPI_ISL_655942\|20 | ···················· | ·A·················· | ····················· | ···················· | ···················· | ··A················· |
| S | 55 hCoV-19/England/ALDP-B5CF2B/2020\|EPI_ISL_655943\|20 | ···················· | ···················· | ····················· | ···················· | ···················· | ···················· |
| S | 56 hCoV-19/England/ALDP-B5C459/2020\|EPI_ISL_655944\|20 | ···················· | ···················· | ····················· | ···················· | ···················· | ···················· |
| S | 57 hCoV-19/England/ALDP-B5C6AE/2020\|EPI_ISL_655946\|20 | ···················· | ···················· | ····················· | ···················· | ···················· | ···················· |
| S | 58 hCoV-19/England/ALDP-B5C32F/2020\|EPI_ISL_655949\|20 | ···················· | ···················· | ····················· | ···················· | ···················· | ···················· |
| S | 59 hCoV-19/England/ALDP-B5C170/2020\|EPI_ISL_655950\|20 | ···················· | ·R·················· | ····················· | ···················· | ···················· | ··R················· |
| S | 60 hCoV-19/England/ALDP-B5C310/2020\|EPI_ISL_655951\|20 | ···················· | ···················· | ····················· | ···················· | ···················· | ···················· |
| S | 61 hCoV-19/England/ALDP-B5CFB2/2020\|EPI_ISL_655952\|20 | ···················· | ···················· | ····················· | ···················· | ···················· | ···················· |
| S | 62 hCoV-19/England/ALDP-B5C161/2020\|EPI_ISL_655953\|20 | ···················· | ···················· | ····················· | ···················· | ···················· | ···················· |
| S | 63 hCoV-19/England/ALDP-B5C4B3/2020\|EPI_ISL_655954\|20 | ···················· | ···················· | ····················· | ···················· | ···················· | ···················· |
| S | 64 hCoV-19/England/ALDP-B5CA57/2020\|EPI_ISL_655955\|20 | ···················· | ···················· | ····················· | ···················· | ···················· | ···················· |
| S | 65 hCoV-19/England/ALDP-B5CCAC/2020\|EPI_ISL_655956\|20 | ···················· | ···················· | ····················· | ···················· | ···················· | ···················· |
| S | 66 hCoV-19/England/QEUH-B66085/2020\|EPI_ISL_655957\|20 | ···················· | ···················· | ····················· | ···················· | ···················· | ···················· |
| S | 67 hCoV-19/England/ALDP-B5C750/2020\|EPI_ISL_655958\|20 | ···················· | ···················· | ····················· | ···················· | ···················· | ···················· |
| S | 68 hCoV-19/England/QEUH-B65FB6/2020\|EPI_ISL_655959\|20 | ···················· | ···················· | ····················· | ···················· | ···················· | ···················· |
| S | 69 hCoV-19/England/ALDP-B5C082/2020\|EPI_ISL_655960\|20 | ···················· | ···················· | ····················· | ···················· | ···················· | ···················· |
| S | 70 hCoV-19/England/ALDP-B5C204/2020\|EPI_ISL_655961\|20 | ···················· | ···················· | ····················· | ···················· | ···················· | ···················· |
| S | 71 hCoV-19/England/ALDP-B5C8E4/2020\|EPI_ISL_655962\|20 | ···················· | ···················· | ····················· | ···················· | ···················· | ···················· |
| S | 72 hCoV-19/England/ALDP-B5BF86/2020\|EPI_ISL_655963\|20 | ···················· | ···················· | ····················· | ···················· | ···················· | ···················· |
| S | 73 hCoV-19/England/ALDP-B5C0DD/2020\|EPI_ISL_655964\|20 | ···················· | ···················· | ····················· | ···················· | ···················· | ···················· |
| S | 74 hCoV-19/England/CAMC-B518EF/2020\|EPI_ISL_658100\|20 | ···················· | ···················· | ····················· | ···················· | ···················· | ···················· |
| S | 75 hCoV-19/England/CAMC-B5471C/2020\|EPI_ISL_658101\|20 | ···················· | ···················· | ····················· | ···················· | ···T················ | ···················· |
| S | 76 hCoV-19/England/MILK-B5A303/2020\|EPI_ISL_658102\|20 | ···················· | ···················· | ····················· | ···················· | ···················· | ···················· |
| S | 77 hCoV-19/England/MILK-B5A576/2020\|EPI_ISL_658103\|20 | ···················· | ···················· | ····················· | ···················· | ···················· | ···················· |
| S | 78 hCoV-19/England/CAMC-B51B7D/2020\|EPI_ISL_658104\|20 | ···················· | ···················· | ····················· | ···················· | ···················· | ···················· |
| S | 79 hCoV-19/England/MILK-B55F31/2020\|EPI_ISL_658105\|20 | ···················· | ···················· | ····················· | ···················· | ···················· | ···················· |
| S | 80 hCoV-19/England/MILK-B5A53A/2020\|EPI_ISL_658106\|20 | ···················· | ···················· | ····················· | ···················· | ···················· | ···················· |
| S | 81 hCoV-19/England/MILK-B5A43D/2020\|EPI_ISL_658107\|20 | ···················· | ···················· | ····················· | ···················· | ···················· | ···················· |
| S | 82 hCoV-19/England/MILK-B55D91/2020\|EPI_ISL_658108\|20 | ···················· | ···················· | ····················· | ···················· | ···················· | ···················· |
| S | 83 hCoV-19/England/MILK-B5A637/2020\|EPI_ISL_658109\|20 | ···················· | ···················· | ····················· | ···················· | ···················· | ···················· |
| S | 84 hCoV-19/England/CAMC-B54828/2020\|EPI_ISL_658110\|20 | ···················· | ···················· | ····················· | ···················· | ···················· | ···················· |
| S | 85 hCoV-19/England/MILK-B55F8C/2020\|EPI_ISL_658111\|20 | ···················· | ···················· | ····················· | ···················· | ···················· | ···················· |
| S | 86 hCoV-19/England/MILK-B55E70/2020\|EPI_ISL_658113\|20 | ···················· | ···················· | ····················· | ···················· | ···················· | ···················· |
| S | 87 hCoV-19/England/MILK-B56194/2020\|EPI_ISL_658117\|20 | ···················· | ···················· | ····················· | ···················· | ···················· | ···················· |
| S | 88 hCoV-19/England/MILK-B55C67/2020\|EPI_ISL_658118\|20 | ···················· | ···················· | ····················· | ···················· | ···················· | ···················· |
| S | 89 hCoV-19/England/CAMC-B51AF8/2020\|EPI_ISL_658119\|20 | ···················· | ···················· | ····················· | ···················· | ···················· | ···················· |
| S | 90 hCoV-19/England/MILK-B56167/2020\|EPI_ISL_658121\|20 | ···················· | ···················· | ····················· | ···················· | ···················· | ···················· |
| S | 91 hCoV-19/England/CAMC-B54925/2020\|EPI_ISL_658122\|20 | ···················· | ···················· | ····················· | ···················· | ···················· | ···················· |
| S | 92 hCoV-19/England/MILK-B5A46A/2020\|EPI_ISL_658123\|20 | ···················· | ···················· | ····················· | ···················· | ···················· | ···················· |
| S | 93 hCoV-19/England/CAMC-B517C4/2020\|EPI_ISL_658124\|20 | ···················· | ···················· | ····················· | ···················· | ···················· | ···················· |
| S | 94 hCoV-19/England/CAMC-B5174C/2020\|EPI_ISL_658125\|20 | ···················· | ···················· | ····················· | ···················· | ·····T·············· | ···················· |
| S | 95 hCoV-19/England/MILK-B5A567/2020\|EPI_ISL_658126\|20 | ···················· | ···················· | ····················· | ···················· | ···················· | ···················· |
| S | 96 hCoV-19/England/MILK-B5A312/2020\|EPI_ISL_658127\|20 | ···················· | ···················· | ····················· | ···················· | ···················· | ···················· |
| S | 97 hCoV-19/England/CAMC-B5180D/2020\|EPI_ISL_658128\|20 | ···················· | ···················· | ····················· | ···················· | ···················· | ···················· |
| S | 98 hCoV-19/England/MILK-B5A3A9/2020\|EPI_ISL_658129\|20 | ···················· | ···················· | ····················· | ···················· | ···················· | ···················· |
| S | 99 hCoV-19/England/MILK-B55DBF/2020\|EPI_ISL_658130\|20 | ···················· | ···················· | ····················· | ···················· | ···················· | ···················· |
| S | 100 hCoV-19/England/CAMC-B518C1/2020\|EPI_ISL_658131\|20 | ···················· | ·A·················· | ····················· | ···················· | ···················· | ··A················· |
| S | 101 hCoV-19/England/MILK-B55DA0/2020\|EPI_ISL_658132\|20 | ···················· | ···················· | ····················· | ···················· | ···················· | ···················· |
| S | 102 hCoV-19/England/CAMC-B545F4/2020\|EPI_ISL_658134\|20 | ···················· | ···················· | ····················· | ···················· | ·····T·············· | ···················· |
| S | 103 hCoV-19/England/MILK-B55E43/2020\|EPI_ISL_658135\|20 | ···················· | ···················· | ····················· | ···················· | ···················· | ···················· |
| S | 104 hCoV-19/England/MILK-B5A691/2020\|EPI_ISL_658136\|20 | ···················· | ···················· | ····················· | ···················· | ·····T·············· | ···················· |
| S | 105 hCoV-19/England/MILK-B55F5F/2020\|EPI_ISL_658137\|20 | ···················· | ···················· | ····················· | ···················· | ···················· | ···················· |
| S | 106 hCoV-19/England/CAMC-B51ABC/2020\|EPI_ISL_658138\|20 | ···················· | ·A·················· | ····················· | ···················· | ···················· | ··A················· |
| S | 107 hCoV-19/England/MILK-B5A85F/2020\|EPI_ISL_658139\|20 | ···················· | ···················· | ····················· | ···················· | ···················· | ···················· |
| S | 108 hCoV-19/England/MILK-B5A3C7/2020\|EPI_ISL_658140\|20 | ···················· | ···················· | ····················· | ···················· | ···················· | ···················· |
| S | 109 hCoV-19/England/MILK-B55DFB/2020\|EPI_ISL_658141\|20 | ···················· | ···················· | ····················· | ···················· | ···················· | ···················· |
| S | 110 hCoV-19/England/CAMC-B544CA/2020\|EPI_ISL_658142\|20 | ···················· | ···················· | ····················· | ···················· | ···················· | ···················· |
| S | 111 hCoV-19/England/MILK-B55EDA/2020\|EPI_ISL_658143\|20 | ···················· | ···················· | ····················· | ···················· | ···················· | ···················· |
| S | 112 hCoV-19/England/MILK-B5A628/2020\|EPI_ISL_658145\|20 | ···················· | ·A·················· | ····················· | ···················· | ···················· | ··A················· |
| S | 113 hCoV-19/England/MILK-B5A35E/2020\|EPI_ISL_658146\|20 | ···················· | ···················· | ····················· | ···················· | ···················· | ···················· |
| S | 114 hCoV-19/England/MILK-B5A743/2020\|EPI_ISL_658147\|20 | ···················· | ···················· | ····················· | ···················· | ···················· | ···················· |
| S | 115 hCoV-19/England/CAMC-B548DD/2020\|EPI_ISL_658148\|20 | ···················· | ·A·················· | ····················· | ···················· | ···················· | ··A················· |
| S | 116 hCoV-19/England/MILK-B561FE/2020\|EPI_ISL_658150\|20 | ···················· | ···················· | ····················· | ···················· | ···················· | ···················· |
| S | 117 hCoV-19/England/MILK-B5A52B/2020\|EPI_ISL_658151\|20 | ···················· | ···················· | ····················· | ···················· | ···················· | ···················· |
| S | 118 hCoV-19/England/MILK-B5A6A0/2020\|EPI_ISL_658152\|20 | ···················· | ·A·················· | ····················· | ···················· | ···················· | ··A················· |
| S | 119 hCoV-19/England/MILK-B5A4F1/2020\|EPI_ISL_658154\|20 | ···················· | ···················· | ····················· | ···················· | ···················· | ···················· |
| S | 120 hCoV-19/England/CAMC-B545A9/2020\|EPI_ISL_658156\|20 | ···················· | ···················· | ····················· | ···················· | ···················· | ···················· |
| S | 121 hCoV-19/England/CAMC-B54873/2020\|EPI_ISL_658157\|20 | ···················· | ···················· | ····················· | ···················· | ···················· | ···················· |
| S | 122 hCoV-19/England/CAMC-B51A52/2020\|EPI_ISL_658159\|20 | ···················· | ···················· | ····················· | ···················· | ···················· | ···················· |
| S | 123 hCoV-19/England/MILK-B5A28E/2020\|EPI_ISL_658180\|20 | ···················· | ···················· | ····················· | ···················· | ···················· | ···················· |
| S | 124 hCoV-19/England/CAMC-B545E5/2020\|EPI_ISL_658181\|20 | ···················· | ···················· | ····················· | ···················· | ···················· | ···················· |
| S | 125 hCoV-19/England/MILK-B5A5FE/2020\|EPI_ISL_658182\|20 | ···················· | ···················· | ····················· | ···················· | ···················· | ···················· |
| S | 126 hCoV-19/England/MILK-B560F1/2020\|EPI_ISL_658183\|20 | ···················· | ···················· | ····················· | ···················· | ···················· | ···················· |
| S | 127 hCoV-19/England/MILK-B55DDD/2020\|EPI_ISL_658184\|20 | ···················· | ···················· | ····················· | ···················· | ···················· | ···················· |

| | | | | | | | |
|---|---|---|---|---|---|---|---|
| S | 128 hCoV-19/England/MILK-B5A488/2020\|EPI_ISL_658185\|20 | ···················· | ···················· | ···················· | ···················· | ···················· | ···················· |
| S | 129 hCoV-19/England/CAMC-B5182B/2020\|EPI_ISL_658186\|20 | ···················· | ···················· | ···················· | ···················· | ···················· | ···················· |
| S | 130 hCoV-19/England/CAMC-B546E2/2020\|EPI_ISL_658187\|20 | ···················· | ···················· | ···················· | ···················· | ···················· | ···················· |
| S | 131 hCoV-19/England/CAMC-B51955/2020\|EPI_ISL_658189\|20 | ···················· | ·A·················· | ···················· | ···················· | ···················· | ··A················· |
| S | 132 hCoV-19/England/CAMC-B51ACB/2020\|EPI_ISL_658190\|20 | ···················· | ···················· | ···················· | ···················· | ···················· | ···················· |
| S | 133 hCoV-19/England/CAMC-B545D6/2020\|EPI_ISL_658191\|20 | ···················· | ···················· | ···················· | ···················· | ···················· | ···················· |
| S | 134 hCoV-19/England/CAMC-B5470D/2020\|EPI_ISL_658192\|20 | ···················· | ···················· | ···················· | ···················· | ···················· | ···················· |
| S | 135 hCoV-19/England/MILK-B5A4A6/2020\|EPI_ISL_658193\|20 | ···················· | ···················· | ···················· | ···················· | ···················· | ···················· |
| S | 136 hCoV-19/England/CAMC-B54749/2020\|EPI_ISL_658194\|20 | ···················· | ···················· | ···················· | ···················· | ···················· | ···················· |
| S | 137 hCoV-19/England/CAMC-B51B6E/2020\|EPI_ISL_658196\|20 | ···················· | ···················· | ···················· | ···················· | ···················· | ···················· |
| S | 138 hCoV-19/England/CAMC-B51ADA/2020\|EPI_ISL_658197\|20 | ···················· | ···················· | ···················· | ···················· | ···················· | ···················· |
| S | 139 hCoV-19/England/CAMC-B54767/2020\|EPI_ISL_658198\|20 | ···················· | ···················· | ···················· | ···················· | ···················· | ···················· |
| S | 140 hCoV-19/England/CAMC-B54776/2020\|EPI_ISL_658199\|20 | ···················· | ···················· | ···················· | ···················· | ···················· | ···················· |
| S | 141 hCoV-19/England/CAMC-B33379/2020\|EPI_ISL_659408\|20 | ···················· | ···················· | ···················· | ···················· | ···················· | ···················· |
| S | 142 hCoV-19/England/CAMC-B3228C/2020\|EPI_ISL_659410\|20 | ···················· | ···················· | ···················· | ···················· | ···················· | ···················· |
| S | 143 hCoV-19/England/CAMC-B33B62/2020\|EPI_ISL_659413\|20 | ···················· | ···················· | ···················· | ···················· | ···················· | ···················· |
| S | 144 hCoV-19/England/CAMC-B330BE/2020\|EPI_ISL_659418\|20 | ···················· | ···················· | ···················· | ···················· | ···················· | ···················· |
| S | 145 hCoV-19/England/CAMC-B32C24/2020\|EPI_ISL_659423\|20 | ···················· | ···················· | ···················· | ···················· | ···················· | ···················· |
| S | 146 hCoV-19/England/CAMC-B3424E/2020\|EPI_ISL_659425\|20 | ···················· | ···················· | ···················· | ···················· | ···················· | ···················· |
| S | 147 hCoV-19/England/CAMC-B354FC/2020\|EPI_ISL_659426\|20 | ···················· | ···················· | ···················· | ···················· | ······T············· | ···················· |
| S | 148 hCoV-19/England/CAMC-B345DC/2020\|EPI_ISL_659427\|20 | ···················· | ···················· | ···················· | ···················· | ···················· | ···················· |
| S | 149 hCoV-19/England/CAMC-B351AA/2020\|EPI_ISL_659428\|20 | ···················· | ·A·················· | ···················· | ···················· | ··········T········· | ···················· |
| S | 150 hCoV-19/England/CAMC-B34141/2020\|EPI_ISL_659429\|20 | ···················· | ·A·················· | ···················· | ···················· | ···················· | ···················· |
| S | 151 hCoV-19/England/CAMC-B34A64/2020\|EPI_ISL_659431\|20 | ···················· | ···················· | ···················· | ···················· | ···················· | ···················· |
| S | 152 hCoV-19/England/CAMC-B355F9/2020\|EPI_ISL_659432\|20 | ···················· | ···················· | ···················· | ···················· | ···················· | ···················· |
| S | 153 hCoV-19/England/CAMC-B34642/2020\|EPI_ISL_659434\|20 | ···················· | ···················· | ···················· | ···················· | ······T············· | ···················· |
| S | 154 hCoV-19/England/CAMC-B343D2/2020\|EPI_ISL_659436\|20 | ···················· | ·A·················· | ···················· | ···················· | ···················· | ··A················· |
| S | 155 hCoV-19/England/CAMC-B34A46/2020\|EPI_ISL_659437\|20 | ···················· | ···················· | ···················· | ···················· | ···················· | ···················· |
| S | 156 hCoV-19/England/CAMC-B34439/2020\|EPI_ISL_659439\|20 | ···················· | ···················· | ···················· | ···················· | ······T············· | ···················· |
| S | 157 hCoV-19/England/CAMC-B35429/2020\|EPI_ISL_659441\|20 | ···················· | ···················· | ···················· | ···················· | ···················· | ···················· |
| S | 158 hCoV-19/England/CAMC-B34484/2020\|EPI_ISL_659443\|20 | ···················· | ···················· | ···················· | ···················· | ···················· | ···················· |
| S | 159 hCoV-19/England/CAMC-B3566F/2020\|EPI_ISL_659444\|20 | ···················· | ···················· | ···················· | ···················· | ···················· | ···················· |
| S | 160 hCoV-19/England/CAMC-B32811/2020\|EPI_ISL_659445\|20 | ···················· | ···················· | ···················· | ···················· | ···················· | ···················· |
| S | 161 hCoV-19/England/CAMC-B32D4F/2020\|EPI_ISL_659447\|20 | ···················· | ···················· | ···················· | ···················· | ···················· | ···················· |
| S | 162 hCoV-19/England/CAMC-B357F3/2020\|EPI_ISL_659448\|20 | ···················· | ···················· | ···················· | ···················· | ···················· | ···················· |
| S | 163 hCoV-19/England/CAMC-B34DF2/2020\|EPI_ISL_659450\|20 | ···················· | ···················· | ···················· | ···················· | ······T············· | ···················· |
| S | 164 hCoV-19/England/CAMC-B32B81/2020\|EPI_ISL_659452\|20 | ···················· | ···················· | ···················· | ···················· | ···················· | ···················· |
| S | 165 hCoV-19/England/CAMC-B328E4/2020\|EPI_ISL_659453\|20 | ···················· | ···················· | ···················· | ···················· | ···················· | ···················· |
| S | 166 hCoV-19/England/CAMC-B35544/2020\|EPI_ISL_659454\|20 | ···················· | ·A·················· | ···················· | ···················· | ···················· | ··A················· |
| S | 167 hCoV-19/England/CAMC-B32969/2020\|EPI_ISL_659455\|20 | ···················· | ···················· | ···················· | ···················· | ···················· | ···················· |
| S | 168 hCoV-19/England/CAMC-B356F6/2020\|EPI_ISL_659456\|20 | ···················· | ···················· | ···················· | ···················· | ···················· | ···················· |
| S | 169 hCoV-19/England/CAMC-B3294B/2020\|EPI_ISL_659457\|20 | ···················· | ···················· | ···················· | ···················· | ···················· | ···················· |
| S | 170 hCoV-19/England/CAMC-B34DB6/2020\|EPI_ISL_659459\|20 | ···················· | ···················· | ···················· | ···················· | ···················· | ···················· |
| S | 171 hCoV-19/England/CAMC-B32B45/2020\|EPI_ISL_659460\|20 | ···················· | ···················· | ···················· | ···················· | ······T············· | ···················· |
| S | 172 hCoV-19/England/CAMC-B34457/2020\|EPI_ISL_659461\|20 | ···················· | ···················· | ···················· | ···················· | ···················· | ···················· |
| S | 173 hCoV-19/England/CAMC-B34ABF/2020\|EPI_ISL_659462\|20 | ···················· | ···················· | ···················· | ···················· | ···················· | ···················· |
| S | 174 hCoV-19/England/CAMC-B32D21/2020\|EPI_ISL_659465\|20 | ···················· | ···················· | ···················· | ···················· | ······T············· | ···················· |
| S | 175 hCoV-19/England/CAMC-B35386/2020\|EPI_ISL_659466\|20 | ···················· | ···················· | ···················· | ···················· | ···················· | ···················· |
| S | 176 hCoV-19/England/CAMC-B357C6/2020\|EPI_ISL_659467\|20 | ···················· | ···················· | ···················· | ···················· | ···················· | ···················· |
| S | 177 hCoV-19/England/CAMC-B3432D/2020\|EPI_ISL_659469\|20 | ···················· | ·A·················· | ···················· | ···················· | ···················· | ··A················· |
| S | 178 hCoV-19/England/CAMC-B3576C/2020\|EPI_ISL_659470\|20 | ···················· | ···················· | ···················· | ···················· | ···················· | ···················· |
| S | 179 hCoV-19/England/CAMC-B346F7/2020\|EPI_ISL_659472\|20 | ···················· | ···················· | ···················· | ···················· | ···················· | ···················· |
| S | 180 hCoV-19/England/CAMC-B350AD/2020\|EPI_ISL_659473\|20 | ···················· | ···················· | ···················· | ···················· | ···················· | ···················· |
| S | 181 hCoV-19/England/CAMC-B32A2A/2020\|EPI_ISL_659474\|20 | ···················· | ···················· | ···················· | ···················· | ···················· | ···················· |
| S | 182 hCoV-19/England/CAMC-B35517/2020\|EPI_ISL_659476\|20 | ···················· | ···················· | ···················· | ···················· | ···················· | ···················· |
| S | 183 hCoV-19/England/CAMC-B3477C/2020\|EPI_ISL_659477\|20 | ···················· | ···················· | ···················· | ···················· | ···················· | ···················· |
| S | 184 hCoV-19/England/CAMC-B3483D/2020\|EPI_ISL_659479\|20 | ···················· | ···················· | ···················· | ···················· | ···················· | ···················· |
| S | 185 hCoV-19/England/CAMC-B32C42/2020\|EPI_ISL_659480\|20 | ···················· | ···················· | ···················· | ···················· | ···················· | ···················· |
| S | 186 hCoV-19/England/CAMC-B35508/2020\|EPI_ISL_659483\|20 | ···················· | ···················· | ···················· | ···················· | ···················· | ···················· |
| S | 187 hCoV-19/England/CAMC-B34712/2020\|EPI_ISL_659485\|20 | ···················· | ···················· | ···················· | ···················· | ···················· | ···················· |
| S | 188 hCoV-19/England/CAMC-B3580F/2020\|EPI_ISL_659486\|20 | ···················· | ···················· | ···················· | ···················· | ···················· | ···················· |
| S | 189 hCoV-19/England/CAMC-B34DA7/2020\|EPI_ISL_659488\|20 | ···················· | ···················· | ···················· | ···················· | ···················· | ···················· |
| S | 190 hCoV-19/England/CAMC-B32AEE/2020\|EPI_ISL_659489\|20 | ···················· | ···················· | ···················· | ···················· | ···················· | ···················· |
| S | 191 hCoV-19/Sweden/20-22304/2020\|EPI_ISL_660400\|2020-0 | ···················· | ···················· | ···················· | ···················· | ···················· | ···················· |
| S | 192 hCoV-19/Sweden/20-22306/2020\|EPI_ISL_660401\|2020-0 | ···················· | ···················· | ···················· | ···················· | ···················· | ···················· |
| S | 193 hCoV-19/Sweden/20-22308/2020\|EPI_ISL_660402\|2020-0 | ···················· | ···················· | ···················· | ···················· | ···················· | ···················· |
| S | 194 hCoV-19/Sweden/20-22309/2020\|EPI_ISL_660403\|2020-0 | ···················· | ···················· | ···················· | ···················· | ···················· | ···················· |
| S | 195 hCoV-19/Sweden/20-22310/2020\|EPI_ISL_660404\|2020-0 | ···················· | ···················· | ···················· | ···················· | ···················· | ···················· |
| S | 196 hCoV-19/Sweden/20-22312/2020\|EPI_ISL_660405\|2020-0 | ···················· | ···················· | ···················· | ···················· | ···················· | ···················· |
| S | 197 hCoV-19/Sweden/20-22313/2020\|EPI_ISL_660406\|2020-0 | ···················· | ···················· | ···················· | ···················· | ···················· | ···················· |
| S | 198 hCoV-19/Sweden/20-22315/2020\|EPI_ISL_660407\|2020-0 | ···················· | ···················· | ···················· | ···················· | ···················· | ···················· |
| S | 199 hCoV-19/Sweden/20-22316/2020\|EPI_ISL_660408\|2020-0 | ···················· | ···················· | ···················· | ···················· | ···················· | ···················· |
| S | 200 hCoV-19/Sweden/20-22317/2020\|EPI_ISL_660409\|2020-0 | ···················· | ···················· | ···················· | ···················· | ···················· | ···················· |
| S | 201 hCoV-19/Sweden/20-22318/2020\|EPI_ISL_660410\|2020-0 | ···················· | ···················· | ···················· | ···················· | ···················· | ···················· |
| S | 202 hCoV-19/Sweden/20-22319/2020\|EPI_ISL_660411\|2020-0 | ···················· | ···················· | ···················· | ···················· | ···················· | ···················· |
| S | 203 hCoV-19/Sweden/20-22321/2020\|EPI_ISL_660412\|2020-0 | ···················· | ···················· | ···················· | ···················· | ···················· | ···················· |
| S | 204 hCoV-19/Sweden/20-22323/2020\|EPI_ISL_660413\|2020-0 | ···················· | ···················· | ···················· | ···················· | ···················· | ···················· |
| S | 205 hCoV-19/Sweden/20-22324/2020\|EPI_ISL_660414\|2020-0 | ···················· | ···················· | ···················· | ···················· | ···················· | ···················· |
| S | 206 hCoV-19/Sweden/20-22561/2020\|EPI_ISL_660415\|2020-0 | ···················· | ···················· | ···················· | ···················· | ···················· | ···················· |
| S | 207 hCoV-19/Sweden/20-52652/2020\|EPI_ISL_660416\|2020-0 | ···················· | ···················· | ···················· | ···················· | ···················· | ···················· |

```
S 208 hCoV-19/Sweden/20-52653/2020|EPI_ISL_660417|2020-0 .................... .................... .................... .................... .................... ....................
S 209 hCoV-19/Sweden/20-52654/2020|EPI_ISL_660418|2020-0 .................... .................... .................... .................... .................... ....................
S 210 hCoV-19/Sweden/20-52655/2020|EPI_ISL_660419|2020-0 .................... .................... .................... .................... .................... ....................
S 211 hCoV-19/Sweden/20-52656/2020|EPI_ISL_660420|2020-0 .................... .................... .................... .................... .................... ....................
S 212 hCoV-19/Sweden/20-52657/2020|EPI_ISL_660421|2020-0 .................... .................... .................... .................... .................... ....................
S 213 hCoV-19/Sweden/20-52931/2020|EPI_ISL_660422|2020-0 .................... .................... .................... .................... .................... ....................
S 214 hCoV-19/Sweden/20-52937/2020|EPI_ISL_660423|2020-0 .................... .................... .................... .................... .................... ....................
S 215 hCoV-19/Sweden/20-52944/2020|EPI_ISL_660424|2020-0 .................... .................... .................... .................... .................... ....................
S 216 hCoV-19/Sweden/20-52951/2020|EPI_ISL_660425|2020-0 .................... .................... .................... .................... .................... ....................
S 217 hCoV-19/Sweden/20-52952/2020|EPI_ISL_660426|2020-0 .................... .................... .................... .................... .................... ....................
S 218 hCoV-19/Sweden/20-52956/2020|EPI_ISL_660427|2020-0 .................... .................... .................... .................... .................... ....................
S 219 hCoV-19/Sweden/20-52958/2020|EPI_ISL_660428|2020-0 .................... .................... .................... .................... .................... ....................
S 220 hCoV-19/Sweden/20-52961/2020|EPI_ISL_660429|2020-0 .................... .................... .................... .................... .................... ....................
S 221 hCoV-19/Sweden/20-52962/2020|EPI_ISL_660430|2020-0 .................... .................... .................... .................... .................... ....................
S 222 hCoV-19/Sweden/20-52963/2020|EPI_ISL_660431|2020-0 .................... .................... .................... .................... .................... ....................
S 223 hCoV-19/France/ARA-SC337/2020|EPI_ISL_660432|2020- .................... .................... .................... .................... .................... ....................
S 224 hCoV-19/Russia/MOS-CRIE-5047186/2020|EPI_ISL_66043 .................... .................... .................... .................... .................... ....................
S 225 hCoV-19/Russia/MOS-CRIE-5047186-2/2020|EPI_ISL_660 .................... .................... .................... .................... .................... ....................
S 226 hCoV-19/Russia/MOS-CRIE-7210452/2020|EPI_ISL_66043 .................... .................... .................... .................... .................... ....................
S 227 hCoV-19/Russia/MOS-CRIE-7161283/2020|EPI_ISL_66043 .................... .................... .................... .................... .................... ....................
S 228 hCoV-19/Burkina Faso/1124/2020|EPI_ISL_660446|2020 .................... .................... .................... .................... .................... ....................
S 229 hCoV-19/Burkina Faso/1447/2020|EPI_ISL_660447|2020 .................... .................... .................... .................... .................... ....................
S 231 hCoV-19/Burkina Faso/1698/2020|EPI_ISL_660449|2020 .................... .................... .................... .................... .................... ....................
S 232 hCoV-19/Burkina Faso/2-009/2020|EPI_ISL_660450|202 .................... .................... .................... .................... .................... ....................
S 233 hCoV-19/Burkina Faso/2-012/2020|EPI_ISL_660451|202 .................... .................... .................... .................... .................... ....................
S 234 hCoV-19/Burkina Faso/2-035-1347/2020|EPI_ISL_66045 .................... .................... .................... .................... .................... ....................
S 235 hCoV-19/Burkina Faso/2-037-1654/2020|EPI_ISL_66045 .................... .................... .................... .................... .................... ....................
S 236 hCoV-19/Burkina Faso/2-039-1610/2020|EPI_ISL_66045 .................... .................... .................... .................... .................... ....................
S 237 hCoV-19/Burkina Faso/2-053-1646/2020|EPI_ISL_66045 .................... .................... .................... .................... .................... ....................
S 238 hCoV-19/Burkina Faso/2-064-2547/2020|EPI_ISL_66045 .................... .................... .................... .................... .................... ....................
S 239 hCoV-19/Burkina Faso/2-068-2228/2020|EPI_ISL_66045 .................... .................... .................... .................... .................... ....................
S 240 hCoV-19/Burkina Faso/2470/2020|EPI_ISL_660458|2020 .................... .................... .................... .................... .................... ....................
S 241 hCoV-19/Burkina Faso/2598/2020|EPI_ISL_660459|2020 .................... .................... .................... .................... .................... ....................
S 242 hCoV-19/Burkina Faso/2636/2020|EPI_ISL_660460|2020 .................... .................... .................... .................... .................... ....................
S 243 hCoV-19/Burkina Faso/2701/2020|EPI_ISL_660461|2020 .................... .................... .................... .................... .................... ....................
S 244 hCoV-19/Burkina Faso/2799/2020|EPI_ISL_660462|2020 .................... .................... .................... .................... .................... ....................
S 245 hCoV-19/Burkina Faso/2832/2020|EPI_ISL_660463|2020 .................... .................... .................... .................... .................... ....................
S 246 hCoV-19/Burkina Faso/2944/2020|EPI_ISL_660464|2020 .................... .................... .................... .................... .................... ....................
S 247 hCoV-19/Burkina Faso/2972/2020|EPI_ISL_660465|2020 .................... .................... .................... .................... .................... ....................
S 248 hCoV-19/Burkina Faso/3052/2020|EPI_ISL_660466|2020 .................... .................... .................... .................... .................... ....................
S 249 hCoV-19/Burkina Faso/3103/2020|EPI_ISL_660467|2020 .................... .................... .................... .................... .................... ....................
S 250 hCoV-19/Burkina Faso/3570/2020|EPI_ISL_660468|2020 .................... .................... .................... .................... .................... ....................
S 251 hCoV-19/Burkina Faso/4053/2020|EPI_ISL_660469|2020 .................... .................... .................... .................... .................... ....................
S 252 hCoV-19/Burkina Faso/4057/2020|EPI_ISL_660470|2020 .................... .................... .................... .................... .................... ....................
S 253 hCoV-19/Burkina Faso/4202/2020|EPI_ISL_660471|2020 .................... .................... .................... .................... .................... ....................
S 254 hCoV-19/Burkina Faso/4215/2020|EPI_ISL_660472|2020 .................... .................... .................... .................... .................... ....................
S 255 hCoV-19/Burkina Faso/4307/2020|EPI_ISL_660473|2020 .................... .................... .................... .................... .................... ....................
S 257 hCoV-19/USA/FL-BPHL-2047/2020|EPI_ISL_653200|2020- .................... .................... .................... .................... .................... ....................
S 258 hCoV-19/USA/FL-BPHL-2050/2020|EPI_ISL_653203|2020- .................... .................... .................... .................... .................... ....................
S 259 hCoV-19/USA/FL-BPHL-2051/2020|EPI_ISL_653204|2020- .................... .................... .................... .................... .................... ....................
S 260 hCoV-19/USA/FL-BPHL-2053/2020|EPI_ISL_653206|2020- .................... .................... .................... .................... .................... ....................
S 261 hCoV-19/USA/FL-BPHL-2054/2020|EPI_ISL_653207|2020- .................... .................... .................... .................... .................... ....................
S 263 hCoV-19/USA/FL-BPHL-2061/2020|EPI_ISL_653214|2020- .................... .................... .................... .................... .................... ....................
S 264 hCoV-19/USA/FL-BPHL-2065/2020|EPI_ISL_653218|2020- .................... .................... .................... .................... .................... ....................
S 265 hCoV-19/USA/FL-BPHL-2067/2020|EPI_ISL_653220|2020- .................... .................... .................... .................... .................... ....................
S 266 hCoV-19/USA/FL-BPHL-2068/2020|EPI_ISL_653221|2020- .................... .................... .................... .................... .................... ....................
S 267 hCoV-19/USA/FL-BPHL-2071/2020|EPI_ISL_653224|2020- .................... .................... .................... .................... .................... ....................
S 268 hCoV-19/USA/FL-BPHL-2073/2020|EPI_ISL_653226|2020- .................... .................... .................... .................... .................... ....................
S 271 hCoV-19/USA/FL-BPHL-2080/2020|EPI_ISL_653233|2020- .................... .................... .................... .................... .................... ....................
S 273 hCoV-19/USA/FL-BPHL-2092/2020|EPI_ISL_653245|2020- .................... .................... .................... .................... .................... ....................
S 274 hCoV-19/USA/FL-BPHL-2100/2020|EPI_ISL_653253|2020- .................... .................... .................... .................... .................... ....................
S 275 hCoV-19/USA/FL-BPHL-2101/2020|EPI_ISL_653254|2020- .................... .................... .................... .................... .................... ....................
S 276 hCoV-19/USA/FL-BPHL-2102/2020|EPI_ISL_653255|2020- .................... .................... .................... .................... .................... ....................
S 277 hCoV-19/USA/FL-BPHL-2103/2020|EPI_ISL_653256|2020- .................... .................... .................... .................... .................... ....................
S 278 hCoV-19/USA/FL-BPHL-2104/2020|EPI_ISL_653257|2020- .................... .................... .................... .................... .................... ....................
S 279 hCoV-19/USA/FL-BPHL-2105/2020|EPI_ISL_653258|2020- .................... .................... .................... .................... .................... ....................
S 280 hCoV-19/USA/FL-BPHL-2106/2020|EPI_ISL_653259|2020- .................... .................... .................... .................... .................... ....................
S 281 hCoV-19/USA/FL-BPHL-2108/2020|EPI_ISL_653261|2020- .................... .................... .................... .................... .................... ....................
S 282 hCoV-19/USA/FL-BPHL-2110/2020|EPI_ISL_653263|2020- .................... .................... .................... .................... .................... ....................
S 283 hCoV-19/USA/FL-BPHL-2112/2020|EPI_ISL_653265|2020- .................... .................... .................... .................... .................... ....................
S 284 hCoV-19/USA/FL-BPHL-2113/2020|EPI_ISL_653266|2020- .................... .................... .................... .................... .................... ....................
S 285 hCoV-19/USA/FL-BPHL-2114/2020|EPI_ISL_653267|2020- .................... .................... .................... .................... .................... ....................
S 286 hCoV-19/USA/FL-BPHL-2115/2020|EPI_ISL_653268|2020- .................... .................... .................... .................... .................... ....................
S 287 hCoV-19/USA/FL-BPHL-2116/2020|EPI_ISL_653269|2020- .................... .................... .................... .................... .................... ....................
S 288 hCoV-19/USA/FL-BPHL-2119/2020|EPI_ISL_653272|2020- .................... .................... .................... .................... .................... ....................
S 289 hCoV-19/USA/FL-BPHL-2120/2020|EPI_ISL_653273|2020- .................... .................... .................... .................... .................... ....................
S 290 hCoV-19/USA/FL-BPHL-2121/2020|EPI_ISL_653274|2020- .................... .................... .................... .................... .................... ....................
S 291 hCoV-19/USA/FL-BPHL-2122/2020|EPI_ISL_653275|2020- .................... .................... .................... .................... .................... ....................
S 292 hCoV-19/USA/FL-BPHL-2123/2020|EPI_ISL_653276|2020- .................... .................... .................... .................... .................... ....................
S 293 hCoV-19/USA/FL-BPHL-2124/2020|EPI_ISL_653277|2020- .................... .................... .................... .................... .................... ....................
```

| | | | | | | | |
|---|---|---|---|---|---|---|---|
| S | 294 hCoV-19/USA/FL-BPHL-2125/2020\|EPI_ISL_653278\|2020- | ···················· | ···················· | ···················· | ···················· | ···················· | ···················· |
| S | 295 hCoV-19/USA/FL-BPHL-2127/2020\|EPI_ISL_653280\|2020- | ···················· | ···················· | ···················· | ···················· | ···················· | ···················· |
| S | 297 hCoV-19/USA/FL-BPHL-2131/2020\|EPI_ISL_653284\|2020- | ···················· | ···················· | ···················· | ···················· | ···················· | ···················· |
| S | 298 hCoV-19/USA/FL-BPHL-2135/2020\|EPI_ISL_653288\|2020- | ···················· | ···················· | ···················· | ···················· | ···················· | ···················· |
| S | 299 hCoV-19/USA/FL-BPHL-2142/2020\|EPI_ISL_653295\|2020- | ···················· | ···················· | ···················· | ···················· | ···················· | ···················· |
| S | 300 hCoV-19/USA/FL-BPHL-2143/2020\|EPI_ISL_653296\|2020- | ···················· | ···················· | ···················· | ···················· | ···················· | ···················· |
| S | 301 hCoV-19/USA/FL-BPHL-2144/2020\|EPI_ISL_653297\|2020- | ···················· | ···················· | ···················· | ···················· | ···················· | ···················· |
| S | 311 hCoV-19/Spain/CT-IBV-97007964/2020\|EPI_ISL_654525\| | ···················· | ···················· | ···················· | ···················· | ···················· | ···················· |
| S | 325 hCoV-19/Spain/PV-IBV-98008454/2020\|EPI_ISL_654547\| | ···················· | ·A·················· | ···················· | ···················· | ···················· | ···················· |
| S | 326 hCoV-19/Spain/PV-IBV-98008265/2020\|EPI_ISL_654553\| | ···················· | ···················· | ···················· | ···················· | ···················· | ···················· |
| S | 327 hCoV-19/Spain/PV-IBV-98008266/2020\|EPI_ISL_654554\| | ···················· | ···················· | ···················· | ···················· | ···················· | ···················· |
| S | 328 hCoV-19/Spain/PV-IBV-98008273/2020\|EPI_ISL_654555\| | ···················· | ···················· | ···················· | ···················· | ···················· | ···················· |
| S | 329 hCoV-19/Spain/PV-IBV-98008274/2020\|EPI_ISL_654556\| | ···················· | ···················· | ···················· | ···················· | ···················· | ···················· |
| S | 330 hCoV-19/Spain/PV-IBV-98008276/2020\|EPI_ISL_654557\| | ···················· | ···················· | ···················· | ···················· | ···················· | ···················· |
| S | 331 hCoV-19/Spain/PV-IBV-98008279/2020\|EPI_ISL_654558\| | ···················· | ···················· | ···················· | ···················· | ···················· | ···················· |
| S | 332 hCoV-19/Spain/PV-IBV-98008281/2020\|EPI_ISL_654559\| | ···················· | ···················· | ···················· | ···················· | ···················· | ···················· |
| S | 333 hCoV-19/Spain/PV-IBV-98008282/2020\|EPI_ISL_654560\| | ···················· | ···················· | ···················· | ···················· | ···················· | ···················· |
| S | 334 hCoV-19/Spain/PV-IBV-98008284/2020\|EPI_ISL_654561\| | ···················· | ···················· | ···················· | ···················· | ···················· | ···················· |
| S | 335 hCoV-19/Spain/PV-IBV-98008298/2020\|EPI_ISL_654562\| | ···················· | ···················· | ···················· | ···················· | ···················· | ···················· |
| S | 336 hCoV-19/Spain/PV-IBV-98008303/2020\|EPI_ISL_654563\| | ···················· | ···················· | ···················· | ···················· | ···················· | ···················· |
| S | 337 hCoV-19/Spain/PV-IBV-98008309/2020\|EPI_ISL_654564\| | ···················· | ···················· | ···················· | ···················· | ···················· | ···················· |
| S | 338 hCoV-19/Spain/PV-IBV-98008313/2020\|EPI_ISL_654565\| | ···················· | ···················· | ···················· | ···················· | ···················· | ···················· |
| S | 339 hCoV-19/Spain/PV-IBV-98008323/2020\|EPI_ISL_654566\| | ···················· | ···················· | ···················· | ···················· | ···················· | ···················· |
| S | 340 hCoV-19/Spain/PV-IBV-98008333/2020\|EPI_ISL_654567\| | ···················· | ···················· | ···················· | ···················· | ···················· | ···················· |
| S | 341 hCoV-19/Spain/PV-IBV-98008335/2020\|EPI_ISL_654568\| | ···················· | ···················· | ···················· | ···················· | ···················· | ···················· |
| S | 342 hCoV-19/Spain/PV-IBV-98008344/2020\|EPI_ISL_654569\| | ···················· | ···················· | ···················· | ···················· | ···················· | ···················· |
| S | 343 hCoV-19/Spain/PV-IBV-98008345/2020\|EPI_ISL_654570\| | ···················· | ·A·················· | ···················· | ···················· | ···················· | ···················· |
| S | 344 hCoV-19/Spain/PV-IBV-98008346/2020\|EPI_ISL_654571\| | ···················· | ···················· | ···················· | ···················· | ···················· | ···················· |
| S | 345 hCoV-19/Spain/PV-IBV-98008353/2020\|EPI_ISL_654572\| | ···················· | ·A·················· | ···················· | ···················· | ···················· | ···················· |
| S | 346 hCoV-19/Spain/PV-IBV-98008356/2020\|EPI_ISL_654573\| | ···················· | ···················· | ···················· | ···················· | ···················· | ···················· |
| S | 347 hCoV-19/Spain/PV-IBV-98008361/2020\|EPI_ISL_654574\| | ···················· | ···················· | ···················· | ···················· | ···················· | ···················· |
| S | 348 hCoV-19/Spain/PV-IBV-98008363/2020\|EPI_ISL_654575\| | ···················· | ···················· | ···················· | ···················· | ···················· | ···················· |
| S | 349 hCoV-19/Spain/PV-IBV-98008364/2020\|EPI_ISL_654576\| | ···················· | ···················· | ···················· | ···················· | ···················· | ···················· |
| S | 350 hCoV-19/Spain/PV-IBV-98008369/2020\|EPI_ISL_654577\| | ···················· | ···················· | ···················· | ···················· | ···················· | ···················· |
| S | 351 hCoV-19/Spain/PV-IBV-98008371/2020\|EPI_ISL_654578\| | ···················· | ···················· | ···················· | ···················· | ···················· | ···················· |
| S | 352 hCoV-19/Spain/PV-IBV-98008378/2020\|EPI_ISL_654579\| | ···················· | ···················· | ···················· | ···················· | ···················· | ···················· |
| S | 353 hCoV-19/Spain/PV-IBV-98008379/2020\|EPI_ISL_654580\| | ···················· | ···················· | ···················· | ···················· | ···················· | ···················· |
| S | 354 hCoV-19/Spain/PV-IBV-98008380/2020\|EPI_ISL_654581\| | ···················· | ···················· | ···················· | ···················· | ···················· | ···················· |
| S | 355 hCoV-19/Spain/PV-IBV-98008385/2020\|EPI_ISL_654582\| | ···················· | ···················· | ···················· | ···················· | ···················· | ···················· |
| S | 356 hCoV-19/Spain/PV-IBV-98008387/2020\|EPI_ISL_654583\| | ···················· | ···················· | ···················· | ···················· | ···················· | ···················· |
| S | 357 hCoV-19/Spain/PV-IBV-98008389/2020\|EPI_ISL_654584\| | ···················· | ···················· | ···················· | ···················· | ···················· | ···················· |
| S | 358 hCoV-19/Spain/PV-IBV-98008391/2020\|EPI_ISL_654585\| | ···················· | ···················· | ···················· | ···················· | ···················· | ···················· |
| S | 359 hCoV-19/Spain/PV-IBV-98008392/2020\|EPI_ISL_654586\| | ···················· | ···················· | ···················· | ···················· | ···················· | ···················· |
| S | 360 hCoV-19/Spain/PV-IBV-98008395/2020\|EPI_ISL_654587\| | ···················· | ···················· | ···················· | ···················· | ···················· | ···················· |
| S | 361 hCoV-19/Spain/PV-IBV-98008399/2020\|EPI_ISL_654588\| | ···················· | ·A·················· | ···················· | ···················· | ···················· | ···················· |
| S | 362 hCoV-19/Spain/PV-IBV-98008404/2020\|EPI_ISL_654589\| | ···················· | ·A·················· | ···················· | ···················· | ···················· | ···················· |
| S | 363 hCoV-19/Spain/PV-IBV-98008407/2020\|EPI_ISL_654590\| | ···················· | ···················· | ···················· | ···················· | ···················· | ···················· |
| S | 364 hCoV-19/Spain/PV-IBV-98008408/2020\|EPI_ISL_654591\| | ···················· | ···················· | ···················· | ···················· | ···················· | ···················· |
| S | 365 hCoV-19/Spain/PV-IBV-98008409/2020\|EPI_ISL_654592\| | ···················· | ·A·················· | ···················· | ···················· | ···················· | ···················· |
| S | 366 hCoV-19/Spain/PV-IBV-98008410/2020\|EPI_ISL_654593\| | ···················· | ·A·················· | ···················· | ···················· | ···················· | ···················· |
| S | 367 hCoV-19/Spain/PV-IBV-98008415/2020\|EPI_ISL_654594\| | ···················· | ···················· | ···················· | ···················· | ···················· | ···················· |
| S | 368 hCoV-19/Spain/PV-IBV-98008417/2020\|EPI_ISL_654595\| | ···················· | ···················· | ···················· | ···················· | ···················· | ···················· |
| S | 369 hCoV-19/Spain/PV-IBV-98008418/2020\|EPI_ISL_654596\| | ···················· | ···················· | ···················· | ···················· | ···················· | ···················· |
| S | 370 hCoV-19/Spain/PV-IBV-98008420/2020\|EPI_ISL_654597\| | ···················· | ···················· | ···················· | ···················· | ···················· | ···················· |
| S | 371 hCoV-19/Spain/PV-IBV-98008422/2020\|EPI_ISL_654598\| | ···················· | ···················· | ···················· | ···················· | ···················· | ···················· |
| S | 372 hCoV-19/Spain/PV-IBV-98008423/2020\|EPI_ISL_654599\| | ···················· | ···················· | ···················· | ···················· | ···················· | ···················· |
| S | 373 hCoV-19/England/ALDP-B5CB27/2020\|EPI_ISL_655844\|20 | ···················· | ···················· | ···················· | ···················· | ···················· | ···················· |
| S | 374 hCoV-19/England/ALDP-B5C592/2020\|EPI_ISL_655845\|20 | ···················· | ···················· | ···················· | ···················· | ···················· | ···················· |
| S | 375 hCoV-19/England/QEUH-B6C7A8/2020\|EPI_ISL_655847\|20 | ···················· | ···················· | ···················· | ···················· | ···················· | ···················· |
| S | 376 hCoV-19/England/ALDP-B5CC60/2020\|EPI_ISL_655849\|20 | ···················· | ···················· | ···················· | ···················· | ···················· | ···················· |
| S | 377 hCoV-19/England/ALDP-B5CA84/2020\|EPI_ISL_655851\|20 | ···················· | ···················· | ···················· | ···················· | ···················· | ···················· |
| S | 378 hCoV-19/England/ALDP-B5BFD1/2020\|EPI_ISL_655852\|20 | ···················· | ···················· | ···················· | ···················· | ···················· | ···················· |
| S | 379 hCoV-19/England/ALDP-B5CE2E/2020\|EPI_ISL_655853\|20 | ···················· | ···················· | ···················· | ···················· | ···················· | ···················· |
| S | 380 hCoV-19/England/QEUH-B660B2/2020\|EPI_ISL_655854\|20 | ···················· | ···················· | ···················· | ···················· | ···················· | ···················· |
| S | 381 hCoV-19/England/ALDP-B5CF0D/2020\|EPI_ISL_655855\|20 | ···················· | ···················· | ···················· | ···················· | ···················· | ···················· |
| S | 382 hCoV-19/England/ALDP-B5C3C5/2020\|EPI_ISL_655857\|20 | ···················· | ···················· | ···················· | ···················· | ···················· | ···················· |
| S | 383 hCoV-19/England/ALDP-B5CCD9/2020\|EPI_ISL_655858\|20 | ···················· | ···················· | ···················· | ···················· | ···················· | ···················· |
| S | 384 hCoV-19/England/ALDP-B5C2C8/2020\|EPI_ISL_655859\|20 | ···················· | ···················· | ···················· | ···················· | ···················· | ···················· |
| S | 385 hCoV-19/England/QEUH-B65EE6/2020\|EPI_ISL_655861\|20 | ···················· | ···················· | ···················· | ···················· | ···················· | ···················· |
| S | 386 hCoV-19/England/ALDP-B5C152/2020\|EPI_ISL_655862\|20 | ···················· | ···················· | ···················· | ···················· | ···················· | ···················· |
| S | 387 hCoV-19/England/ALDP-B5BEA7/2020\|EPI_ISL_655863\|20 | ···················· | ···················· | ···················· | ···················· | ···················· | ···················· |
| S | 388 hCoV-19/England/QEUH-B65E40/2020\|EPI_ISL_655868\|20 | ···················· | ···················· | ···················· | ···················· | ···················· | ···················· |
| S | 389 hCoV-19/England/ALDP-B5CA1B/2020\|EPI_ISL_655869\|20 | ···················· | ···················· | ···················· | ···················· | ···················· | ···················· |
| S | 390 hCoV-19/England/ALDP-B5C222/2020\|EPI_ISL_655870\|20 | ···················· | ···················· | ···················· | ···················· | ···················· | ···················· |
| S | 391 hCoV-19/England/ALDP-B5C1F8/2020\|EPI_ISL_655871\|20 | ···················· | ···················· | ···················· | ···················· | ···················· | ···················· |
| S | 392 hCoV-19/England/ALDP-B5BF68/2020\|EPI_ISL_655872\|20 | ···················· | ···················· | ···················· | ···················· | ···················· | ···················· |
| S | 393 hCoV-19/England/QEUH-B65F4D/2020\|EPI_ISL_655874\|20 | ···················· | ···················· | ···················· | ···················· | ···················· | ···················· |
| S | 394 hCoV-19/England/ALDP-B5C143/2020\|EPI_ISL_655875\|20 | ···················· | ·A·················· | ···················· | ···················· | ···················· | ···A················ |
| S | 395 hCoV-19/England/ALDP-B5C1AD/2020\|EPI_ISL_655876\|20 | ···················· | ···················· | ···················· | ···················· | ···················· | ···················· |
| S | 396 hCoV-19/England/ALDP-B5C583/2020\|EPI_ISL_655877\|20 | ···················· | ···················· | ···················· | ···················· | ···················· | ···················· |

S 397 hCoV-19/England/ALDP-B5BCCB/2020|EPI_ISL_655878|20 ···················· ···················· ····················· ···················· ·····T·············· ····················
S 398 hCoV-19/England/QEUH-B65F98/2020|EPI_ISL_655879|20 ···················· ···················· ····················· ···················· ···················· ····················
S 399 hCoV-19/England/ALDP-B5C091/2020|EPI_ISL_655880|20 ···················· ···················· ····················· ···················· ···················· ····················
S 400 hCoV-19/England/QEUH-B65EB9/2020|EPI_ISL_655883|20 ···················· ···················· ····················· ···················· ···················· ····················
S 401 hCoV-19/England/ALDP-B5C9B4/2020|EPI_ISL_655884|20 ···················· ···················· ····················· ···················· ···················· ····················
S 402 hCoV-19/England/ALDP-B5BD9B/2020|EPI_ISL_655885|20 ···················· ···················· ····················· ···················· ···················· ····················
S 403 hCoV-19/England/ALDP-B5C723/2020|EPI_ISL_655887|20 ···················· ···················· ····················· ···················· ·····T·············· ····················
S 404 hCoV-19/England/ALDP-B5C028/2020|EPI_ISL_655888|20 ···················· ···················· ····················· ···················· ···················· ····················
S 405 hCoV-19/England/ALDP-B5C996/2020|EPI_ISL_655889|20 ···················· ···················· ····················· ···················· ···················· ····················
S 406 hCoV-19/England/ALDP-B5CEF1/2020|EPI_ISL_655890|20 ···················· ···················· ····················· ···················· ···················· ····················
S 407 hCoV-19/England/QEUH-B65EAA/2020|EPI_ISL_655891|20 ···················· ···················· ····················· ···················· ···················· ····················
S 408 hCoV-19/England/ALDP-B5C34D/2020|EPI_ISL_655892|20 ···················· ···················· ····················· ···················· ···················· ··A·················
S 409 hCoV-19/England/ALDP-B5C107/2020|EPI_ISL_655893|20 ···················· ·A·················· ····················· ···················· ···················· ··A·················
S 410 hCoV-19/England/ALDP-B5CE79/2020|EPI_ISL_655894|20 ···················· ···················· ····················· ···················· ···················· ····················
S 411 hCoV-19/England/ALDP-B5BFB3/2020|EPI_ISL_655895|20 ···················· ···················· ····················· ···················· ···················· ····················
S 412 hCoV-19/England/ALDP-B5C055/2020|EPI_ISL_655896|20 ···················· ···················· ····················· ···················· ···················· ····················
S 413 hCoV-19/England/ALDP-B5C565/2020|EPI_ISL_655897|20 ···················· ···················· ····················· ···················· ···················· ····················
S 414 hCoV-19/England/ALDP-B5CE88/2020|EPI_ISL_655898|20 ···················· ···················· ····················· ···················· ···················· ····················
S 415 hCoV-19/England/ALDP-B5CBCD/2020|EPI_ISL_655899|20 ···················· ···················· ····················· ···················· ···················· ····················
S 416 hCoV-19/England/CAMC-B5167C/2020|EPI_ISL_658020|20 ···················· ···················· ····················· ···················· ···················· ····················
S 417 hCoV-19/England/MILK-B54AAA/2020|EPI_ISL_658021|20 ···················· ···················· ····················· ···················· ···················· ····················
S 418 hCoV-19/England/MILK-B552ED/2020|EPI_ISL_658022|20 ···················· ···················· ····················· ···················· ···················· ····················
S 419 hCoV-19/England/MILK-B54BA7/2020|EPI_ISL_658023|20 ···················· ···················· ····················· ···················· ···················· ····················
S 420 hCoV-19/England/CAMC-B5114E/2020|EPI_ISL_658024|20 ···················· ···················· ····················· ···················· ···················· ····················
S 421 hCoV-19/England/MILK-B5507A/2020|EPI_ISL_658025|20 ···················· ···················· ····················· ···················· ···················· ····················
S 422 hCoV-19/England/MILK-B54A31/2020|EPI_ISL_658026|20 ···················· ···················· ····················· ···················· ···················· ····················
S 423 hCoV-19/England/MILK-B54A22/2020|EPI_ISL_658027|20 ···················· ···················· ····················· ···················· ···················· ····················
S 424 hCoV-19/England/CAMC-B51DC2/2020|EPI_ISL_658028|20 ···················· ···················· ····················· ···················· ···················· ····················
S 425 hCoV-19/England/CAMC-B51111/2020|EPI_ISL_658029|20 ···················· ···················· ····················· ···················· ···················· ····················
S 426 hCoV-19/England/CAMC-B5205F/2020|EPI_ISL_658031|20 ···················· ···················· ····················· ···················· ···················· ····················
S 427 hCoV-19/England/CAMC-B51C2F/2020|EPI_ISL_658032|20 ···················· ···················· ····················· ···················· ···················· ····················
S 428 hCoV-19/England/CAMC-B51C3E/2020|EPI_ISL_658033|20 ···················· ···················· ····················· ···················· ·····T·············· ····················
S 429 hCoV-19/England/CAMC-B514DC/2020|EPI_ISL_658034|20 ···················· ···················· ····················· ···················· ···················· ····················
S 430 hCoV-19/England/MILK-B5504D/2020|EPI_ISL_658035|20 ···················· ···················· ····················· ···················· ···················· ····················
S 431 hCoV-19/England/CAMC-B511F3/2020|EPI_ISL_658036|20 ···················· ···················· ····················· ···················· ···················· ····················
S 432 hCoV-19/England/CAMC-B510D8/2020|EPI_ISL_658037|20 ···················· ···················· ····················· ···················· ···················· ····················
S 433 hCoV-19/England/CAMC-B5472B/2020|EPI_ISL_658160|20 ···················· ···················· ····················· ···················· ···················· ····················
S 434 hCoV-19/England/CAMC-B54679/2020|EPI_ISL_658161|20 ···················· ···················· ····················· ···················· ···················· ····················
S 435 hCoV-19/England/CAMC-B54943/2020|EPI_ISL_658162|20 ···················· ···················· ····················· ···················· ···················· ····················
S 436 hCoV-19/England/CAMC-B5457C/2020|EPI_ISL_658164|20 ···················· ···················· ····················· ···················· ···················· ····················
S 437 hCoV-19/England/CAMC-B515CA/2020|EPI_ISL_658166|20 ···················· ·A·················· ····················· ···················· ···················· ··A·················
S 438 hCoV-19/England/CAMC-B51630/2020|EPI_ISL_658167|20 ···················· ···················· ····················· ···················· ···················· ····················
S 439 hCoV-19/England/MILK-B5A655/2020|EPI_ISL_658168|20 ···················· ···················· ····················· ···················· ···················· ····················
S 440 hCoV-19/England/MILK-B5A27F/2020|EPI_ISL_658169|20 ···················· ···················· ····················· ···················· ···················· ····················
S 441 hCoV-19/England/MILK-B5A2BB/2020|EPI_ISL_658170|20 ···················· ···················· ····················· ···················· ···················· ····················
S 442 hCoV-19/England/CAMC-B518FE/2020|EPI_ISL_658171|20 ···················· ···················· ····················· ···················· ···················· ····················
S 443 hCoV-19/England/MILK-B55F40/2020|EPI_ISL_658172|20 ···················· ···················· ····················· ···················· ···················· ····················
S 444 hCoV-19/England/MILK-B56097/2020|EPI_ISL_658173|20 ···················· ···················· ····················· ···················· ···················· ····················
S 445 hCoV-19/England/MILK-B55D37/2020|EPI_ISL_658174|20 ···················· ···················· ····················· ···················· ···················· ····················
S 446 hCoV-19/England/MILK-B5A45B/2020|EPI_ISL_658176|20 ···················· ···················· ····················· ···················· ···················· ····················
S 447 hCoV-19/England/CAMC-B51982/2020|EPI_ISL_658177|20 ···················· ···················· ····················· ···················· ···················· ····················
S 448 hCoV-19/England/CAMC-B51CA7/2020|EPI_ISL_658178|20 ···················· ···················· ····················· ···················· ···················· ····················
S 449 hCoV-19/England/CAMC-B517A6/2020|EPI_ISL_658179|20 ···················· ···················· ····················· ···················· ···················· ····················
S 450 hCoV-19/England/CAMC-B3342B/2020|EPI_ISL_659351|20 ···················· ·A·················· ····················· ···················· ···················· ··A·················
S 451 hCoV-19/England/CAMC-B335A0/2020|EPI_ISL_659352|20 ···················· ·A·················· ····················· ···················· ···················· ··A·················
S 452 hCoV-19/England/CAMC-B33607/2020|EPI_ISL_659353|20 ···················· ···················· ····················· ···················· ···················· ····················
S 453 hCoV-19/England/CAMC-B33485/2020|EPI_ISL_659354|20 ···················· ···················· ····················· ···················· ···················· ····················
S 454 hCoV-19/England/CAMC-B330FA/2020|EPI_ISL_659355|20 ···················· ···················· ····················· ···················· ···················· ····················
S 455 hCoV-19/England/CAMC-B33018/2020|EPI_ISL_659357|20 ···················· ···················· ····················· ···················· ···················· ····················
S 456 hCoV-19/England/CAMC-B3340D/2020|EPI_ISL_659358|20 ···················· ···················· ····················· ···················· ···················· ····················
S 457 hCoV-19/England/CAMC-B335CE/2020|EPI_ISL_659359|20 ···················· ···················· ····················· ···················· ···················· ····················
S 458 hCoV-19/England/CAMC-B33A65/2020|EPI_ISL_659361|20 ···················· ···················· ····················· ···················· ···················· ····················
S 459 hCoV-19/England/CAMC-B325ED/2020|EPI_ISL_659362|20 ···················· ···················· ····················· ···················· ·····T·············· ····················
S 460 hCoV-19/England/CAMC-B32F58/2020|EPI_ISL_659363|20 ···················· ···················· ····················· ···················· ·····T·············· ····················
S 461 hCoV-19/England/CAMC-B32E6A/2020|EPI_ISL_659364|20 ···················· ···················· ····················· ···················· ···················· ····················
S 462 hCoV-19/England/CAMC-B32547/2020|EPI_ISL_659367|20 ···················· ···················· ····················· ···················· ·····T·············· ····················
S 463 hCoV-19/England/CAMC-B33CBA/2020|EPI_ISL_659368|20 ···················· ···················· ····················· ···················· ···················· ····················
S 464 hCoV-19/England/CAMC-B32FA3/2020|EPI_ISL_659401|20 ···················· ···················· ····················· ···················· ···················· ····················
S 465 hCoV-19/England/CAMC-B33722/2020|EPI_ISL_659402|20 ···················· ···················· ····················· ···················· ···················· ····················
S 466 hCoV-19/England/CAMC-B33F66/2020|EPI_ISL_659403|20 ···················· ·A·················· ····················· ···················· ···················· ····················
S 467 hCoV-19/England/CAMC-B3385C/2020|EPI_ISL_659404|20 ···················· ···················· ····················· ···················· ···················· ····················
S 468 hCoV-19/England/CAMC-B3350A/2020|EPI_ISL_659405|20 ···················· ·A·················· ····················· ···················· ···················· ··A·················
S 469 hCoV-19/England/CAMC-B32486/2020|EPI_ISL_659406|20 ···················· ···················· ····················· ···················· ···················· ····················
S 470 hCoV-19/England/CAMC-B32574/2020|EPI_ISL_659407|20 ···················· ···················· ····················· ···················· ···················· ····················
S 471 hCoV-19/England/CAMC-B32978/2020|EPI_ISL_659490|20 ···················· ···················· ····················· ···················· ···················· ····················
S 472 hCoV-19/England/CAMC-B3479A/2020|EPI_ISL_659492|20 ···················· ···················· ····················· ···················· ···················· ····················
S 473 hCoV-19/England/CAMC-B32DB8/2020|EPI_ISL_659493|20 ···················· ···················· ····················· ···················· ···················· ····················
S 474 hCoV-19/England/CAMC-B3476D/2020|EPI_ISL_659495|20 ···················· ···················· ····················· ···················· ···················· ····················
S 475 hCoV-19/England/CAMC-B34ADD/2020|EPI_ISL_659496|20 ···················· ···················· ····················· ···················· ···················· ····················
S 476 hCoV-19/England/CAMC-B348C4/2020|EPI_ISL_659498|20 ···················· ···················· ····················· ···················· ···················· ····················

| | | | | | | | |
|---|---|---|---|---|---|---|---|
| S | 477 hCoV-19/France/ARA-SC328/2020\|EPI_ISL_660332\|2020- | ···················· | ···················· | ···················· | ···················· | ···················· | ···················· |
| S | 478 hCoV-19/France/ARA-SC331/2020\|EPI_ISL_660333\|2020- | ···················· | ···················· | ···················· | ···················· | ···················· | ···················· |
| S | 479 hCoV-19/France/ARA-SC338/2020\|EPI_ISL_660334\|2020- | ···················· | ···················· | ···················· | ···················· | ···················· | ···················· |
| S | 480 hCoV-19/France/ARA-SC339/2020\|EPI_ISL_660335\|2020- | ···················· | ···················· | ···················· | ···················· | ···················· | ···················· |
| S | 481 hCoV-19/France/ARA-SC341/2020\|EPI_ISL_660336\|2020- | ···················· | ···················· | ···················· | ···················· | ···················· | ···················· |
| S | 482 hCoV-19/France/ARA-SC342/2020\|EPI_ISL_660337\|2020- | ···················· | ···················· | ···················· | ···················· | ···················· | ···················· |
| S | 483 hCoV-19/France/ARA-SC343/2020\|EPI_ISL_660338\|2020- | ···················· | ···················· | ···················· | ···················· | ···················· | ···················· |
| S | 484 hCoV-19/France/ARA-SC344/2020\|EPI_ISL_660339\|2020- | ···················· | ···················· | ···················· | ···················· | ···················· | ···················· |
| S | 485 hCoV-19/France/ARA-SC345/2020\|EPI_ISL_660340\|2020- | ···················· | ···················· | ···················· | ···················· | ···················· | ···················· |
| S | 486 hCoV-19/France/ARA-SC348/2020\|EPI_ISL_660341\|2020- | ···················· | ···················· | ···················· | ···················· | ···················· | ···················· |
| S | 487 hCoV-19/France/ARA-SC349/2020\|EPI_ISL_660342\|2020- | ···················· | ···················· | ···················· | ···················· | ···················· | ···················· |
| S | 488 hCoV-19/France/ARA-SC368/2020\|EPI_ISL_660350\|2020- | ···················· | ···················· | ···················· | ···················· | ···················· | ···················· |
| S | 489 hCoV-19/France/ARA-SC370/2020\|EPI_ISL_660351\|2020- | ···················· | ···················· | ···················· | ···················· | ···················· | ···················· |
| S | 490 hCoV-19/France/ARA-SC371/2020\|EPI_ISL_660352\|2020- | ···················· | ···················· | ···················· | ···················· | ···················· | ···················· |
| S | 491 hCoV-19/France/ARA-SC378/2020\|EPI_ISL_660353\|2020- | ···················· | ···················· | ···················· | ···················· | ···················· | ···················· |
| S | 492 hCoV-19/Burkina Faso/588/2020\|EPI_ISL_660475\|2020- | ···················· | ···················· | ···················· | ···················· | ···················· | ···················· |
| S | 493 hCoV-19/Burkina Faso/590/2020\|EPI_ISL_660476\|2020- | ···················· | ···················· | ···················· | ···················· | ···················· | ···················· |
| S | 494 hCoV-19/Burkina Faso/608/2020\|EPI_ISL_660477\|2020- | ···················· | ···················· | ···················· | ···················· | ···················· | ···················· |
| S | 495 hCoV-19/Burkina Faso/1174/2020\|EPI_ISL_660478\|2020 | ···················· | ···················· | ···················· | ···················· | ···················· | ···················· |
| S | 497 hCoV-19/Burkina Faso/2307/2020\|EPI_ISL_660480\|2020 | ···················· | ···················· | ···················· | ···················· | ···················· | ···················· |
| S | 498 hCoV-19/Burkina Faso/2359/2020\|EPI_ISL_660481\|2020 | ···················· | ···················· | ···················· | ···················· | ···················· | ···················· |
| S | 499 hCoV-19/Burkina Faso/2366/2020\|EPI_ISL_660482\|2020 | ···················· | ···················· | ···················· | ···················· | ···················· | ···················· |
| S | 500 hCoV-19/Burkina Faso/2439/2020\|EPI_ISL_660483\|2020 | ···················· | ···················· | ···················· | ···················· | ···················· | ···················· |
| S | 501 hCoV-19/Burkina Faso/2446/2020\|EPI_ISL_660484\|2020 | ···················· | ···················· | ···················· | ···················· | ···················· | ···················· |
| S | 503 hCoV-19/Burkina Faso/2473/2020\|EPI_ISL_660486\|2020 | ···················· | ···················· | ···················· | ···················· | ···················· | ···················· |
| S | 504 hCoV-19/Burkina Faso/2496/2020\|EPI_ISL_660487\|2020 | ···················· | ···················· | ···················· | ···················· | ···················· | ···················· |
| S | 505 hCoV-19/Burkina Faso/3567/2020\|EPI_ISL_660488\|2020 | ···················· | ···················· | ···················· | ···················· | ···················· | ···················· |
| S | 506 hCoV-19/Burkina Faso/3952/2020\|EPI_ISL_660489\|2020 | ···················· | ···················· | ···················· | ···················· | ···················· | ···················· |
| S | 507 hCoV-19/Burkina Faso/2-040-1615/2020\|EPI_ISL_66049 | ···················· | ···················· | ···················· | ···················· | ···················· | ···················· |
| S | 508 hCoV-19/Burkina Faso/2342/2020\|EPI_ISL_660491\|2020 | ···················· | ···················· | ···················· | ···················· | ···················· | ···················· |
| S | 509 hCoV-19/Burkina Faso/2-041-1649/2020\|EPI_ISL_66049 | ···················· | ···················· | ···················· | ···················· | ···················· | ···················· |
| S | 510 hCoV-19/Burkina Faso/2-042-1650/2020\|EPI_ISL_66049 | ···················· | ···················· | ···················· | ···················· | ···················· | ···················· |
| S | 511 hCoV-19/Burkina Faso/2667/2020\|EPI_ISL_660494\|2020 | ···················· | ···················· | ···················· | ···················· | ···················· | ···················· |
| S | 512 hCoV-19/Burkina Faso/3970/2020\|EPI_ISL_660495\|2020 | ···················· | ···················· | ···················· | ···················· | ···················· | ···················· |
| S | 513 hCoV-19/France/NAQ-SC222/2020\|EPI_ISL_649970\|2020- | ···················· | ···················· | ···················· | ···················· | ···················· | ···················· |
| S | 514 hCoV-19/France/NAQ-SC223/2020\|EPI_ISL_649971\|2020- | ···················· | ···················· | ···················· | ···················· | ···················· | ···················· |
| S | 515 hCoV-19/USA/MI-UM-10036596128/2020\|EPI_ISL_649973\| | ···················· | ···················· | ···················· | ···················· | ···················· | ···················· |
| S | 516 hCoV-19/USA/MI-UM-10036605682/2020\|EPI_ISL_649975\| | ···················· | ···················· | ···················· | ···················· | ···················· | ···················· |
| S | 518 hCoV-19/USA/MI-UM-10036609489/2020\|EPI_ISL_649979\| | ···················· | ·A·················· | ···················· | ···················· | ···················· | ···················· |
| S | 520 hCoV-19/USA/MI-UM-10036599544/2020\|EPI_ISL_649982\| | ···················· | ···················· | ···················· | ···················· | ···················· | ···················· |
| S | 523 hCoV-19/USA/MI-UM-10036593588/2020\|EPI_ISL_649985\| | ···················· | ···················· | ···················· | ···················· | ···················· | ···················· |
| S | 524 hCoV-19/USA/MI-UM-10036589205/2020\|EPI_ISL_649987\| | ···················· | ···················· | ···················· | ···················· | ···················· | ···················· |
| S | 528 hCoV-19/USA/MI-UM-10036587103/2020\|EPI_ISL_649991\| | ···················· | ···················· | ···················· | ···················· | ···················· | ···················· |
| S | 530 hCoV-19/USA/MI-UM-10036587309/2020\|EPI_ISL_649993\| | ···················· | ···················· | ···················· | ···················· | ···················· | ···················· |
| S | 531 hCoV-19/USA/MI-UM-10036595910/2020\|EPI_ISL_649994\| | ···················· | ·A·················· | ···················· | ···················· | ···················· | ···················· |
| S | 532 hCoV-19/USA/MI-UM-10036597152/2020\|EPI_ISL_649996\| | ···················· | ···················· | ···················· | ···················· | ···················· | ···················· |
| S | 533 hCoV-19/USA/MI-UM-10036579461/2020\|EPI_ISL_649998\| | ···················· | ···················· | ···················· | ···················· | ···················· | ···················· |
| S | 534 hCoV-19/USA/MI-UM-10036740029/2020\|EPI_ISL_649999\| | ···················· | ······A············· | ···················· | ···················· | ···················· | ···················· |
| S | 535 hCoV-19/Sweden/20-23815/2020\|EPI_ISL_654500\|2020-1 | ···················· | ···················· | ···················· | ···················· | ···················· | ···················· |
| S | 536 hCoV-19/Sweden/20-23816/2020\|EPI_ISL_654501\|2020-1 | ···················· | ···················· | ···················· | ···················· | ···················· | ···················· |
| S | 537 hCoV-19/Sweden/20-53268/2020\|EPI_ISL_654502\|2020-1 | ···················· | ···················· | ···················· | ···················· | ···················· | ···················· |
| S | 538 hCoV-19/Sweden/20-53266/2020\|EPI_ISL_654503\|2020-1 | ···················· | ···················· | ···················· | ···················· | ···················· | ···················· |
| S | 539 hCoV-19/Sweden/20-53044/2020\|EPI_ISL_654504\|2020-1 | ···················· | ·A·················· | ···················· | ···················· | ···················· | ···A················ |
| S | 540 hCoV-19/Sweden/20-53045/2020\|EPI_ISL_654505\|2020-1 | ···················· | ·A·················· | ···················· | ···················· | ···················· | ···A················ |
| S | 541 hCoV-19/Sweden/20-52706/2020\|EPI_ISL_654507\|2020-0 | ···················· | ···················· | ···················· | ···················· | ···················· | ···················· |
| S | 542 hCoV-19/Sweden/20-23680/2020\|EPI_ISL_654508\|2020-1 | ···················· | ···················· | ···················· | ···················· | ···················· | ···················· |
| S | 543 hCoV-19/England/ALDP-B5CE3D/2020\|EPI_ISL_655800\|20 | ···················· | ···················· | ···················· | ···················· | ···················· | ···················· |
| S | 544 hCoV-19/England/ALDP-B5C50B/2020\|EPI_ISL_655801\|20 | ···················· | ···················· | ···················· | ···················· | ···················· | ···················· |
| S | 545 hCoV-19/England/ALDP-B5C29B/2020\|EPI_ISL_655802\|20 | ···················· | ···················· | ···················· | ···················· | ···················· | ···················· |
| S | 546 hCoV-19/England/ALDP-B5CCBB/2020\|EPI_ISL_655803\|20 | ···················· | ···················· | ···················· | ···················· | ···················· | ···················· |
| S | 547 hCoV-19/England/QEUH-B678D7/2020\|EPI_ISL_655804\|20 | ···················· | ···················· | ···················· | ···················· | ···················· | ···················· |
| S | 548 hCoV-19/England/ALDP-B5CEC4/2020\|EPI_ISL_655805\|20 | ···················· | ·········T·········· | ···················· | ···················· | ···················· | ···················· |
| S | 549 hCoV-19/England/QEUH-B65FC5/2020\|EPI_ISL_655806\|20 | ···················· | ···················· | ···················· | ···················· | ···················· | ···················· |
| S | 550 hCoV-19/England/QEUH-B67901/2020\|EPI_ISL_655807\|20 | ···················· | ···················· | ···················· | ···················· | ···················· | ···················· |
| S | 551 hCoV-19/England/ALDP-B5C9C3/2020\|EPI_ISL_655808\|20 | ···················· | ···················· | ···················· | ···················· | ···················· | ···················· |
| S | 552 hCoV-19/England/QEUH-B6C720/2020\|EPI_ISL_655809\|20 | ········T··········· | ···················· | ···················· | ···················· | ···················· | ···················· |
| S | 553 hCoV-19/England/ALDP-B5CF76/2020\|EPI_ISL_655811\|20 | ···················· | ···················· | ···················· | ···················· | ···················· | ···················· |
| S | 554 hCoV-19/England/ALDP-B5CB54/2020\|EPI_ISL_655812\|20 | ···················· | ···················· | ···················· | ···················· | ···················· | ···················· |
| S | 555 hCoV-19/England/ALDP-B5C8D5/2020\|EPI_ISL_655813\|20 | ···················· | ···················· | ···················· | ···················· | ···················· | ···················· |
| S | 556 hCoV-19/England/ALDP-B5CBAF/2020\|EPI_ISL_655815\|20 | ···················· | ···················· | ···················· | ···················· | ···················· | ···················· |
| S | 557 hCoV-19/England/ALDP-B5C969/2020\|EPI_ISL_655816\|20 | ···················· | ···················· | ···················· | ···················· | ···················· | ···················· |
| S | 558 hCoV-19/England/QEUH-B6C6BA/2020\|EPI_ISL_655817\|20 | ···················· | ···················· | ···················· | ···················· | ···················· | ···················· |
| S | 559 hCoV-19/England/QEUH-B6C6E7/2020\|EPI_ISL_655819\|20 | ···················· | ···················· | ···················· | ···················· | ···················· | ···················· |
| S | 560 hCoV-19/England/ALDP-B5CAB1/2020\|EPI_ISL_655820\|20 | ···················· | ···················· | ···················· | ···················· | ···················· | ···················· |
| S | 561 hCoV-19/England/ALDP-B5C2E6/2020\|EPI_ISL_655821\|20 | ···················· | ···················· | ···················· | ···················· | ···················· | ···················· |
| S | 562 hCoV-19/England/ALDP-B5BF2C/2020\|EPI_ISL_655823\|20 | ···················· | ···················· | ···················· | ···················· | ···················· | ···················· |
| S | 563 hCoV-19/England/ALDP-B5C820/2020\|EPI_ISL_655824\|20 | ···················· | ···················· | ···················· | ···················· | ···················· | ···················· |
| S | 564 hCoV-19/England/ALDP-B5C78D/2020\|EPI_ISL_655827\|20 | ···················· | ···················· | ···················· | ···················· | ···················· | ···················· |
| S | 565 hCoV-19/England/ALDP-B5C714/2020\|EPI_ISL_655829\|20 | ···················· | ···················· | ···················· | ···················· | ···················· | ···················· |
| S | 566 hCoV-19/England/ALDP-B5C4E0/2020\|EPI_ISL_655830\|20 | ···················· | ·A·················· | ···················· | ···················· | ···················· | ···················· |

| | | | | | | | |
|---|---|---|---|---|---|---|---|
| S | 567 hCoV-19/England/ALDP-B5CB18/2020\|EPI_ISL_655831\|20 | ···················· | ···················· | ····················· | ···················· | ···················· | ···················· |
| S | 568 hCoV-19/England/ALDP-B5C3A7/2020\|EPI_ISL_655835\|20 | ···················· | ···················· | ····················· | ···················· | ···················· | ···················· |
| S | 569 hCoV-19/England/ALDP-B5BE3E/2020\|EPI_ISL_655836\|20 | ···················· | ···················· | ····················· | ···················· | ···················· | ···················· |
| S | 570 hCoV-19/England/ALDP-B5BED4/2020\|EPI_ISL_655837\|20 | ···················· | ···················· | ····················· | ···················· | ···················· | ···················· |
| S | 571 hCoV-19/England/ALDP-B5CE00/2020\|EPI_ISL_655838\|20 | ···················· | ···················· | ····················· | ···················· | ···················· | ···················· |
| S | 572 hCoV-19/England/ALDP-B5BE4D/2020\|EPI_ISL_655839\|20 | ···················· | ···················· | ····················· | ···················· | ···················· | ···················· |
| S | 573 hCoV-19/England/ALDP-B5C5CF/2020\|EPI_ISL_655840\|20 | ···················· | ···················· | ····················· | ···················· | ···················· | ···················· |
| S | 574 hCoV-19/England/QEUH-B6C799/2020\|EPI_ISL_655841\|20 | ···················· | ···················· | ····················· | ···················· | ···················· | ···················· |
| S | 575 hCoV-19/England/ALDP-B5C240/2020\|EPI_ISL_655842\|20 | ···················· | ···················· | ····················· | ···················· | ···················· | ···················· |
| S | 576 hCoV-19/England/CAMC-B51E0B/2020\|EPI_ISL_658001\|20 | ···················· | ···················· | ····················· | ···················· | ···················· | ···················· |
| S | 577 hCoV-19/England/MILK-B553EA/2020\|EPI_ISL_658002\|20 | ···················· | ···················· | ····················· | ···················· | ···················· | ···················· |
| S | 578 hCoV-19/England/MILK-B5570C/2020\|EPI_ISL_658003\|20 | ···················· | ···················· | ····················· | ···················· | ···················· | ···················· |
| S | 579 hCoV-19/England/MILK-B554AB/2020\|EPI_ISL_658004\|20 | ···················· | ···················· | ····················· | ···················· | ···················· | ···················· |
| S | 580 hCoV-19/England/MILK-B553DB/2020\|EPI_ISL_658005\|20 | ···················· | ·A·················· | ····················· | ···················· | ···················· | ···A················ |
| S | 581 hCoV-19/England/MILK-B54B01/2020\|EPI_ISL_658006\|20 | ···················· | ···················· | ····················· | ···················· | ···················· | ···················· |
| S | 582 hCoV-19/England/CAMC-B521A7/2020\|EPI_ISL_658007\|20 | ···················· | ···················· | ····················· | ···················· | ···················· | ···················· |
| S | 583 hCoV-19/England/CAMC-B51CE3/2020\|EPI_ISL_658008\|20 | ···················· | ···················· | ····················· | ···················· | ···················· | ···················· |
| S | 584 hCoV-19/England/MILK-B551B3/2020\|EPI_ISL_658009\|20 | ···················· | ···················· | ····················· | ···················· | ···················· | ···················· |
| S | 585 hCoV-19/England/CAMC-B51D2C/2020\|EPI_ISL_658010\|20 | ···················· | ···················· | ····················· | ···················· | ···················· | ···················· |
| S | 586 hCoV-19/England/CAMC-B51C5C/2020\|EPI_ISL_658011\|20 | ···················· | ···················· | ····················· | ···················· | ···················· | ···················· |
| S | 587 hCoV-19/England/CAMC-B5215C/2020\|EPI_ISL_658012\|20 | ···················· | ···················· | ····················· | ···················· | ···················· | ···················· |
| S | 588 hCoV-19/England/CAMC-B51436/2020\|EPI_ISL_658013\|20 | ···················· | ···················· | ····················· | ···················· | ···················· | ···················· |
| S | 589 hCoV-19/England/MILK-B55308/2020\|EPI_ISL_658015\|20 | ···················· | ···················· | ····················· | ···················· | ···················· | ···················· |
| S | 590 hCoV-19/England/MILK-B555B7/2020\|EPI_ISL_658016\|20 | ···················· | ···················· | ····················· | ···················· | ···················· | ···················· |
| S | 591 hCoV-19/England/MILK-B54F50/2020\|EPI_ISL_658017\|20 | ···················· | ···················· | ····················· | ···················· | ···················· | ···················· |
| S | 592 hCoV-19/England/MILK-B553F9/2020\|EPI_ISL_658018\|20 | ···················· | ···················· | ····················· | ···················· | ···················· | ···················· |
| S | 593 hCoV-19/England/CAMC-B51D1D/2020\|EPI_ISL_658019\|20 | ···················· | ···················· | ····················· | ···················· | ···················· | ···················· |
| S | 594 hCoV-19/England/MILK-B54C86/2020\|EPI_ISL_658038\|20 | ···················· | ···················· | ····················· | ···················· | ···················· | ···················· |
| S | 595 hCoV-19/England/MILK-B55186/2020\|EPI_ISL_658039\|20 | ···················· | ···················· | ····················· | ···················· | ···················· | ···················· |
| S | 596 hCoV-19/England/MILK-B54EDB/2020\|EPI_ISL_658040\|20 | ···················· | ···················· | ····················· | ···················· | ···················· | ···················· |
| S | 597 hCoV-19/England/CAMC-B51F80/2020\|EPI_ISL_658041\|20 | ···················· | ···················· | ····················· | ···················· | ···················· | ···················· |
| S | 598 hCoV-19/England/MILK-B54D92/2020\|EPI_ISL_658042\|20 | ···················· | ···················· | ····················· | ···················· | ···················· | ···················· |
| S | 599 hCoV-19/England/CAMC-B5206E/2020\|EPI_ISL_658043\|20 | ···················· | ···················· | ····················· | ···················· | ···················· | ···················· |
| S | 600 hCoV-19/England/MILK-B54A40/2020\|EPI_ISL_658044\|20 | ···················· | ···················· | ····················· | ···················· | ···················· | ···················· |
| S | 601 hCoV-19/England/CAMC-B5107E/2020\|EPI_ISL_658045\|20 | ···················· | ···················· | ····················· | ···················· | ···················· | ···················· |
| S | 602 hCoV-19/England/CAMC-B51102/2020\|EPI_ISL_658046\|20 | ···················· | ···················· | ····················· | ···················· | ···················· | ···················· |
| S | 603 hCoV-19/England/MILK-B55353/2020\|EPI_ISL_658047\|20 | ···················· | ···················· | ····················· | ···················· | ···················· | ···················· |
| S | 604 hCoV-19/England/CAMC-B512A5/2020\|EPI_ISL_658048\|20 | ···················· | ···················· | ····················· | ···················· | ···················· | ···················· |
| S | 605 hCoV-19/England/MILK-B55177/2020\|EPI_ISL_658049\|20 | ···················· | ···················· | ····················· | ···················· | ···················· | ···················· |
| S | 606 hCoV-19/England/MILK-B55450/2020\|EPI_ISL_658050\|20 | ···················· | ·A·················· | ····················· | ···················· | ···················· | ···A················ |
| S | 607 hCoV-19/England/MILK-B54B4D/2020\|EPI_ISL_658051\|20 | ···················· | ···················· | ····················· | ···················· | ···················· | ···················· |
| S | 608 hCoV-19/England/CAMC-B51409/2020\|EPI_ISL_658052\|20 | ···················· | ···················· | ····················· | ···················· | ···················· | ···················· |
| S | 609 hCoV-19/England/MILK-B54A9B/2020\|EPI_ISL_658053\|20 | ···················· | ···················· | ····················· | ···················· | ···················· | ···················· |
| S | 610 hCoV-19/England/CAMC-B51603/2020\|EPI_ISL_658055\|20 | ···················· | ···················· | ····················· | ···················· | ···················· | ···················· |
| S | 611 hCoV-19/England/CAMC-B513EE/2020\|EPI_ISL_658056\|20 | ···················· | ···················· | ····················· | ···················· | ···················· | ···················· |
| S | 612 hCoV-19/England/MILK-B54CE0/2020\|EPI_ISL_658057\|20 | ···················· | ···················· | ····················· | ···················· | ···················· | ···················· |
| S | 613 hCoV-19/England/MILK-B54E08/2020\|EPI_ISL_658060\|20 | ···················· | ···················· | ····················· | ···················· | ···················· | ···················· |
| S | 614 hCoV-19/England/MILK-B5A673/2020\|EPI_ISL_658061\|20 | ···················· | ···················· | ····················· | ···················· | ···················· | ···················· |
| S | 615 hCoV-19/England/CAMC-B5169A/2020\|EPI_ISL_658063\|20 | ···················· | ···················· | ····················· | ···················· | ···················· | ···················· |
| S | 616 hCoV-19/England/MILK-B55EAD/2020\|EPI_ISL_658065\|20 | ···················· | ···················· | ····················· | ···················· | ···················· | ···················· |
| S | 617 hCoV-19/England/MILK-B55BA6/2020\|EPI_ISL_658066\|20 | ···················· | ···················· | ····················· | ···················· | ···················· | ···················· |
| S | 618 hCoV-19/England/MILK-B5A44C/2020\|EPI_ISL_658067\|20 | ···················· | ···················· | ····················· | ···················· | ···················· | ···················· |
| S | 619 hCoV-19/England/MILK-B55BD3/2020\|EPI_ISL_658070\|20 | ···················· | ···················· | ····················· | ···················· | ···················· | ···················· |
| S | 620 hCoV-19/England/CAMC-B519FB/2020\|EPI_ISL_658071\|20 | ···················· | ···················· | ····················· | ···················· | ···················· | ···················· |
| S | 621 hCoV-19/England/CAMC-B516C7/2020\|EPI_ISL_658073\|20 | ···················· | ···················· | ····················· | ···················· | ···················· | ···················· |
| S | 622 hCoV-19/England/CAMC-B51928/2020\|EPI_ISL_658074\|20 | ···················· | ···················· | ····················· | ···················· | ···················· | ···················· |
| S | 623 hCoV-19/England/CAMC-B547D0/2020\|EPI_ISL_658075\|20 | ···················· | ···················· | ····················· | ···················· | ···················· | ···················· |
| S | 624 hCoV-19/England/CAMC-B548CE/2020\|EPI_ISL_658076\|20 | ···················· | ···················· | ····················· | ···················· | ···················· | ···················· |
| S | 625 hCoV-19/England/MILK-B5610D/2020\|EPI_ISL_658079\|20 | ···················· | ·A·················· | ····················· | ···················· | ···················· | ···················· |
| S | 626 hCoV-19/England/CAMC-B517E2/2020\|EPI_ISL_658080\|20 | ···················· | ···················· | ····················· | ···················· | ···················· | ···················· |
| S | 627 hCoV-19/England/MILK-B5A42E/2020\|EPI_ISL_658081\|20 | ···················· | ···················· | ····················· | ···················· | ···················· | ···················· |
| S | 628 hCoV-19/England/MILK-B55C76/2020\|EPI_ISL_658082\|20 | ···················· | ···················· | ····················· | ···················· | ···················· | ···················· |
| S | 629 hCoV-19/England/CAMC-B54600/2020\|EPI_ISL_658084\|20 | ···················· | ···················· | ····················· | ···················· | ···················· | ···················· |
| S | 630 hCoV-19/England/MILK-B55CC1/2020\|EPI_ISL_658087\|20 | ···················· | ···················· | ····················· | ···················· | ···················· | ···················· |
| S | 631 hCoV-19/England/MILK-B5A2F7/2020\|EPI_ISL_658088\|20 | ···················· | ···················· | ····················· | ···················· | ···················· | ···················· |
| S | 632 hCoV-19/England/MILK-B55F22/2020\|EPI_ISL_658090\|20 | ···················· | ·A·················· | ····················· | ···················· | ···················· | ···A················ |
| S | 633 hCoV-19/England/CAMC-B5172E/2020\|EPI_ISL_658093\|20 | ···················· | ·A·················· | ····················· | ···················· | ···················· | ···A················ |
| S | 634 hCoV-19/England/MILK-B55D28/2020\|EPI_ISL_658094\|20 | ···················· | ···················· | ····················· | ···················· | ···················· | ···················· |
| S | 635 hCoV-19/England/CAMC-B545C7/2020\|EPI_ISL_658095\|20 | ···················· | ···················· | ····················· | ···················· | ······T············· | ···················· |
| S | 636 hCoV-19/England/CAMC-B5473A/2020\|EPI_ISL_658097\|20 | ···················· | ···················· | ····················· | ···················· | ···················· | ···················· |
| S | 637 hCoV-19/England/CAMC-B5454F/2020\|EPI_ISL_658099\|20 | ···················· | ···················· | ····················· | ···················· | ···················· | ···················· |
| S | 638 hCoV-19/England/CAMC-B3341C/2020\|EPI_ISL_659301\|20 | ···················· | ···················· | ····················· | ···················· | ···················· | ···················· |
| S | 639 hCoV-19/England/CAMC-B3387A/2020\|EPI_ISL_659303\|20 | ···················· | ···················· | ····················· | ···················· | ···················· | ···················· |
| S | 640 hCoV-19/England/CAMC-B32E79/2020\|EPI_ISL_659304\|20 | ···················· | ···················· | ····················· | ···················· | ···················· | ···················· |
| S | 641 hCoV-19/England/CAMC-B33B26/2020\|EPI_ISL_659306\|20 | ···················· | ···················· | ····················· | ···················· | ···················· | ···················· |
| S | 642 hCoV-19/England/CAMC-B32E5B/2020\|EPI_ISL_659307\|20 | ···················· | ···················· | ····················· | ···················· | ···················· | ···················· |
| S | 643 hCoV-19/England/CAMC-B32680/2020\|EPI_ISL_659309\|20 | ···················· | ···················· | ····················· | ···················· | ···················· | ···················· |
| S | 644 hCoV-19/England/CAMC-B32F3A/2020\|EPI_ISL_659311\|20 | ···················· | ·A·················· | ····················· | ···················· | ···················· | ···A················ |
| S | 645 hCoV-19/England/CAMC-B3382F/2020\|EPI_ISL_659312\|20 | ···················· | ···················· | ····················· | ···················· | ···················· | ···················· |
| S | 646 hCoV-19/England/CAMC-B32F76/2020\|EPI_ISL_659314\|20 | ···················· | ···················· | ····················· | ···················· | ···················· | ···················· |

S 647 hCoV-19/England/CAMC-B3343A/2020|EPI_ISL_659315|20 ···················· ···················· ····················· ···················· ······T············· ····················
S 648 hCoV-19/England/CAMC-B33E1E/2020|EPI_ISL_659316|20 ···················· ···················· ····················· ···················· ···················· ····················
S 649 hCoV-19/England/CAMC-B33D02/2020|EPI_ISL_659317|20 ···················· ···················· ····················· ···················· ···················· ····················
S 650 hCoV-19/England/CAMC-B33C6F/2020|EPI_ISL_659319|20 ···················· ···················· ····················· ···················· ···················· ····················
S 651 hCoV-19/England/CAMC-B33072/2020|EPI_ISL_659321|20 ···················· ···················· ····················· ···················· ···················· ····················
S 652 hCoV-19/England/CAMC-B33519/2020|EPI_ISL_659323|20 ···················· ···················· ····················· ···················· ······T············· ····················
S 653 hCoV-19/England/CAMC-B33C9C/2020|EPI_ISL_659326|20 ···················· ···················· ····················· ···················· ···················· ····················
S 654 hCoV-19/England/CAMC-B33449/2020|EPI_ISL_659327|20 ···················· ···················· ····················· ···················· ···················· ····················
S 655 hCoV-19/England/CAMC-B32C15/2020|EPI_ISL_659328|20 ···················· ···················· ····················· ···················· ···················· ····················
S 656 hCoV-19/England/CAMC-B33A92/2020|EPI_ISL_659331|20 ···················· ···················· ····················· ···················· ···················· ····················
S 657 hCoV-19/England/CAMC-B33977/2020|EPI_ISL_659332|20 ···················· ···················· ····················· ···················· ···················· ····················
S 658 hCoV-19/England/CAMC-B3326D/2020|EPI_ISL_659334|20 ···················· ···················· ····················· ···················· ···················· ····················
S 659 hCoV-19/England/CAMC-B33106/2020|EPI_ISL_659335|20 ···················· ···················· ····················· ···················· ···················· ····················
S 660 hCoV-19/England/CAMC-B3391D/2020|EPI_ISL_659336|20 ···················· ···················· ····················· ···················· ···················· ····················
S 661 hCoV-19/England/CAMC-B335FB/2020|EPI_ISL_659337|20 ···················· ···················· ····················· ···················· ···················· ····················
S 662 hCoV-19/England/CAMC-B33142/2020|EPI_ISL_659338|20 ···················· ···················· ····················· ···················· ···················· ····················
S 663 hCoV-19/England/CAMC-B334EF/2020|EPI_ISL_659339|20 ···················· ···················· ····················· ···················· ···················· ····················
S 664 hCoV-19/England/CAMC-B338E3/2020|EPI_ISL_659340|20 ···················· ···················· ····················· ···················· ···················· ····················
S 665 hCoV-19/England/CAMC-B32BAF/2020|EPI_ISL_659342|20 ···················· ···················· ····················· ···················· ···················· ····················
S 666 hCoV-19/England/CAMC-B32213/2020|EPI_ISL_659344|20 ···················· ···················· ····················· ···················· ···················· ···A················
S 667 hCoV-19/England/CAMC-B33CC9/2020|EPI_ISL_659346|20 ···················· ···················· ····················· ···················· ···················· ····················
S 668 hCoV-19/England/CAMC-B32E3D/2020|EPI_ISL_659347|20 ···················· ···················· ····················· ···················· ···················· ····················
S 669 hCoV-19/England/CAMC-B330AF/2020|EPI_ISL_659369|20 ···················· ···················· ····················· ···················· ···················· ····················
S 670 hCoV-19/England/CAMC-B32BDC/2020|EPI_ISL_659370|20 ···················· ···················· ····················· ···················· ···················· ····················
S 671 hCoV-19/England/CAMC-B33889/2020|EPI_ISL_659371|20 ···················· ·A·················· ····················· ···················· ···················· ···A················
S 672 hCoV-19/England/CAMC-B33F84/2020|EPI_ISL_659374|20 ···················· ···················· ····················· ···················· ···················· ····················
S 673 hCoV-19/England/CAMC-B33CAB/2020|EPI_ISL_659375|20 ···················· ···················· ····················· ···················· ···················· ····················
S 674 hCoV-19/England/CAMC-B33F57/2020|EPI_ISL_659376|20 ···················· ···················· ····················· ···················· ···················· ····················
S 675 hCoV-19/England/CAMC-B32FEF/2020|EPI_ISL_659377|20 ···················· ···················· ····················· ···················· ···················· ····················
S 676 hCoV-19/England/CAMC-B3237A/2020|EPI_ISL_659379|20 ···················· ···················· ····················· ···················· ···················· ····················
S 677 hCoV-19/England/CAMC-B33528/2020|EPI_ISL_659381|20 ···················· ···················· ····················· ···················· ···················· ····················
S 678 hCoV-19/England/CAMC-B32204/2020|EPI_ISL_659382|20 ···················· ···················· ····················· ···················· ···················· ····················
S 679 hCoV-19/England/CAMC-B322D7/2020|EPI_ISL_659383|20 ···················· ···················· ····················· ···················· ···················· ····················
S 680 hCoV-19/England/CAMC-B32608/2020|EPI_ISL_659384|20 ···················· ···················· ····················· ···················· ···················· ····················
S 681 hCoV-19/England/CAMC-B323C5/2020|EPI_ISL_659385|20 ···················· ···················· ····················· ···················· ···················· ····················
S 682 hCoV-19/England/CAMC-B3368F/2020|EPI_ISL_659387|20 ···················· ···················· ····················· ···················· ···················· ····················
S 683 hCoV-19/England/CAMC-B33C7E/2020|EPI_ISL_659388|20 ···················· ···················· ····················· ···················· ···················· ····················
S 684 hCoV-19/England/CAMC-B32538/2020|EPI_ISL_659391|20 ···················· ···················· ····················· ···················· ···················· ····················
S 685 hCoV-19/England/CAMC-B32BEB/2020|EPI_ISL_659392|20 ···················· ···················· ····················· ···················· ···················· ····················
S 686 hCoV-19/England/CAMC-B33458/2020|EPI_ISL_659393|20 ···················· ·A·················· ····················· ···················· ···················· ···A················
S 687 hCoV-19/England/CAMC-B33E4B/2020|EPI_ISL_659394|20 ···················· ···················· ····················· ···················· ···················· ····················
S 688 hCoV-19/England/CAMC-B3242C/2020|EPI_ISL_659398|20 ···················· ···················· ····················· ···················· ···················· ····················
S 689 hCoV-19/England/CAMC-B32671/2020|EPI_ISL_659399|20 ···················· ···················· ····················· ···················· ···················· ····················
S 704 hCoV-19/Spain/CT-IBV-99009917/2020|EPI_ISL_660314| ···················· ···················· ····················· ···················· ···················· ····················
S 712 hCoV-19/France/CVL-IPP9550/2020|EPI_ISL_660323|202 ···················· ···················· ····················· ···················· ···················· ····················
S 713 hCoV-19/France/CVL-IPP9554/2020|EPI_ISL_660324|202 ···················· ···················· ····················· ···················· ···················· ····················
S 714 hCoV-19/France/IDF-IPP9555/2020|EPI_ISL_660325|202 ···················· ···················· ····················· ···················· ···················· ····················
S 715 hCoV-19/France/ARA-SC304/2020|EPI_ISL_660326|2020- ···················· ···················· ····················· ···················· ···················· ····················
S 716 hCoV-19/France/ARA-SC321/2020|EPI_ISL_660327|2020- ···················· ···················· ····················· ···················· ···················· ····················
S 717 hCoV-19/France/ARA-SC323/2020|EPI_ISL_660328|2020- ···················· ···················· ····················· ···················· ···················· ····················
S 718 hCoV-19/France/ARA-SC325/2020|EPI_ISL_660329|2020- ···················· ···················· ····················· ···················· ···················· ····················
S 719 hCoV-19/France/ARA-SC326/2020|EPI_ISL_660330|2020- ···················· ···················· ····················· ···················· ···················· ····················
S 720 hCoV-19/France/ARA-SC327/2020|EPI_ISL_660331|2020- ···················· ···················· ····················· ···················· ···················· ····················
S 721 hCoV-19/France/ARA-SC352/2020|EPI_ISL_660343|2020- ···················· ···················· ····················· ···················· ···················· ····················
S 722 hCoV-19/France/ARA-SC358/2020|EPI_ISL_660344|2020- ···················· ···················· ····················· ···················· ···················· ····················
S 723 hCoV-19/France/ARA-SC360/2020|EPI_ISL_660345|2020- ···················· ···················· ····················· ···················· ···················· ····················
S 724 hCoV-19/France/ARA-SC361/2020|EPI_ISL_660346|2020- ···················· ···················· ····················· ···················· ···················· ····················
S 725 hCoV-19/France/ARA-SC362/2020|EPI_ISL_660347|2020- ···················· ···················· ····················· ···················· ···················· ····················
S 726 hCoV-19/France/ARA-SC363/2020|EPI_ISL_660348|2020- ···················· ···················· ····················· ···················· ···················· ····················
S 727 hCoV-19/France/ARA-SC365/2020|EPI_ISL_660349|2020- ···················· ···················· ····················· ···················· ···················· ····················
S 728 hCoV-19/France/ARA-SC381/2020|EPI_ISL_660354|2020- ···················· ···················· ····················· ···················· ···················· ····················
S 729 hCoV-19/France/ARA-SC382/2020|EPI_ISL_660355|2020- ···················· ···················· ····················· ···················· ···················· ····················
S 730 hCoV-19/France/ARA-SC384/2020|EPI_ISL_660356|2020- ···················· ···················· ····················· ···················· ···················· ····················
S 731 hCoV-19/France/ARA-SC385/2020|EPI_ISL_660357|2020- ···················· ···················· ····················· ···················· ···················· ····················
S 732 hCoV-19/France/ARA-SC386/2020|EPI_ISL_660358|2020- ···················· ···················· ····················· ···················· ···················· ····················
S 733 hCoV-19/France/ARA-SC389/2020|EPI_ISL_660359|2020- ···················· ···················· ····················· ···················· ···················· ····················
S 734 hCoV-19/France/ARA-SC391/2020|EPI_ISL_660360|2020- ···················· ···················· ····················· ···················· ···················· ····················
S 735 hCoV-19/France/ARA-SC396/2020|EPI_ISL_660361|2020- ···················· ···················· ····················· ···················· ···················· ····················
S 736 hCoV-19/France/ARA-SC399/2020|EPI_ISL_660362|2020- ···················· ···················· ····················· ···················· ···················· ····················
S 737 hCoV-19/France/ARA-SC400/2020|EPI_ISL_660363|2020- ···················· ···················· ····················· ···················· ···················· ····················
S 738 hCoV-19/France/ARA-SC402/2020|EPI_ISL_660364|2020- ···················· ···················· ····················· ···················· ···················· ····················
S 739 hCoV-19/France/ARA-SC411/2020|EPI_ISL_660370|2020- ···················· ···················· ···········T········· ···················· ···················· ····················
S 740 hCoV-19/France/ARA-SC414/2020|EPI_ISL_660371|2020- ···················· ···················· ···········T········· ···················· ···················· ····················
S 741 hCoV-19/France/NAQ-SC226/2020|EPI_ISL_660372|2020- ···················· ···················· ····················· ···················· ···················· ····················
S 742 hCoV-19/France/OCC-SC419/2020|EPI_ISL_660373|2020- ···················· ···················· ····················· ···················· ···················· ····················
S 743 hCoV-19/France/OCC-SC420/2020|EPI_ISL_660374|2020- ···················· ···················· ····················· ···················· ···················· ····················
S 744 hCoV-19/France/OCC-SC422/2020|EPI_ISL_660375|2020- ···················· ···················· ····················· ···················· ···················· ····················
S 745 hCoV-19/France/OCC-SC423/2020|EPI_ISL_660376|2020- ···················· ···················· ····················· ···················· ···················· ····················
S 746 hCoV-19/France/OCC-SC424/2020|EPI_ISL_660377|2020- ···················· ···················· ····················· ···················· ···················· ····················
S 747 hCoV-19/Sweden/20-22127/2020|EPI_ISL_660378|2020-0 ···················· ···················· ····················· ···················· ···················· ····················

| | | | | | | | |
|---|---|---|---|---|---|---|---|
| S | 748 hCoV-19/Sweden/20-22130/2020\|EPI_ISL_660379\|2020-0 | .................... | .................... | ..................... | .................... | .................... | .................... |
| S | 749 hCoV-19/Sweden/20-22132/2020\|EPI_ISL_660380\|2020-0 | .................... | .................... | ..................... | .................... | .................... | .................... |
| S | 750 hCoV-19/Sweden/20-22133/2020\|EPI_ISL_660381\|2020-0 | .................... | .................... | ..................... | .................... | .................... | .................... |
| S | 751 hCoV-19/Sweden/20-22134/2020\|EPI_ISL_660382\|2020-0 | .................... | .................... | ..................... | .................... | .................... | .................... |
| S | 752 hCoV-19/Sweden/20-22135/2020\|EPI_ISL_660383\|2020-0 | .................... | .................... | ..................... | .................... | .................... | .................... |
| S | 753 hCoV-19/Sweden/20-22286/2020\|EPI_ISL_660384\|2020-0 | .................... | .................... | ..................... | .................... | .................... | .................... |
| S | 754 hCoV-19/Sweden/20-22287/2020\|EPI_ISL_660385\|2020-0 | .................... | .................... | ..................... | .................... | .................... | .................... |
| S | 755 hCoV-19/Sweden/20-22288/2020\|EPI_ISL_660386\|2020-0 | .................... | .................... | ..................... | .................... | .................... | .................... |
| S | 756 hCoV-19/Sweden/20-22289/2020\|EPI_ISL_660387\|2020-0 | .................... | .................... | ..................... | .................... | .................... | .................... |
| S | 757 hCoV-19/Sweden/20-22291/2020\|EPI_ISL_660388\|2020-0 | .................... | .................... | ..................... | .................... | .................... | .................... |
| S | 758 hCoV-19/Sweden/20-22292/2020\|EPI_ISL_660389\|2020-0 | .................... | .................... | ..................... | .................... | .................... | .................... |
| S | 759 hCoV-19/Sweden/20-22293/2020\|EPI_ISL_660390\|2020-0 | .................... | .................... | ..................... | .................... | .................... | .................... |
| S | 760 hCoV-19/Sweden/20-22294/2020\|EPI_ISL_660391\|2020-0 | .................... | .................... | ..................... | .................... | .................... | .................... |
| S | 761 hCoV-19/Sweden/20-22295/2020\|EPI_ISL_660392\|2020-0 | .................... | .................... | ..................... | .................... | .................... | .................... |
| S | 762 hCoV-19/Sweden/20-22296/2020\|EPI_ISL_660393\|2020-0 | .................... | .................... | ..................... | .................... | .................... | .................... |
| S | 763 hCoV-19/Sweden/20-22297/2020\|EPI_ISL_660394\|2020-0 | .................... | .................... | ..................... | .................... | .................... | .................... |
| S | 764 hCoV-19/Sweden/20-22298/2020\|EPI_ISL_660395\|2020-0 | .................... | .................... | ..................... | .................... | .................... | .................... |
| S | 765 hCoV-19/Sweden/20-22299/2020\|EPI_ISL_660396\|2020-0 | .................... | .................... | ..................... | .................... | .................... | .................... |
| S | 766 hCoV-19/Sweden/20-22300/2020\|EPI_ISL_660397\|2020-0 | .................... | .................... | ..................... | .................... | .................... | .................... |
| S | 767 hCoV-19/Sweden/20-22301/2020\|EPI_ISL_660398\|2020-0 | .................... | .................... | ..................... | .................... | .................... | .................... |
| S | 768 hCoV-19/Sweden/20-22302/2020\|EPI_ISL_660399\|2020-0 | .................... | .................... | ..................... | .................... | .................... | .................... |
| S | 769 hCoV-19/England/204700086/2020\|EPI_ISL_649890\|2020 | .................... | .................... | ..................... | .................... | .................... | .................... |
| S | 770 hCoV-19/England/204720609/2020\|EPI_ISL_649891\|2020 | .................... | .................... | ..................... | .................... | .................... | .................... |
| S | 771 hCoV-19/England/204742224/2020\|EPI_ISL_649927\|2020 | .................... | .................... | ..................... | .................... | .................... | .................... |
| S | 772 hCoV-19/England/204742481/2020\|EPI_ISL_649936\|2020 | .................... | .................... | ..................... | .................... | .................... | .................... |
| S | 773 hCoV-19/England/204742482/2020\|EPI_ISL_649937\|2020 | .................... | .................... | ..................... | .................... | .................... | .................... |
| S | 774 hCoV-19/Italy/APU-IZSPB-190PT/2020\|EPI_ISL_649938\| | .................... | .................... | ..................... | .................... | .................... | .................... |
| S | 775 hCoV-19/Italy/APU-IZSPB-192PT/2020\|EPI_ISL_649939\| | .................... | .................... | ..................... | .................... | .................... | .................... |
| S | 776 hCoV-19/Italy/APU-IZSPB-193APT/2020\|EPI_ISL_649940 | .................... | .................... | ..................... | .................... | .................... | .................... |
| S | 777 hCoV-19/France/ARA-SC303/2020\|EPI_ISL_649942\|2020- | .................... | .................... | ..................... | .................... | .................... | .................... |
| S | 778 hCoV-19/France/ARA-SC305/2020\|EPI_ISL_649943\|2020- | .................... | .................... | ..................... | .................... | .................... | .................... |
| S | 779 hCoV-19/France/ARA-SC306/2020\|EPI_ISL_649944\|2020- | .................... | .................... | ..................... | .................... | .................... | .................... |
| S | 780 hCoV-19/France/ARA-SC307/2020\|EPI_ISL_649945\|2020- | .................... | .................... | ..................... | .................... | .................... | .................... |
| S | 781 hCoV-19/France/ARA-SC308/2020\|EPI_ISL_649946\|2020- | .................... | .................... | ..................... | .................... | .................... | .................... |
| S | 782 hCoV-19/France/ARA-SC312/2020\|EPI_ISL_649947\|2020- | .................... | .................... | ..................... | .................... | .................... | .................... |
| S | 783 hCoV-19/France/ARA-SC313/2020\|EPI_ISL_649948\|2020- | .................... | .................... | ..................... | .................... | .................... | .................... |
| S | 784 hCoV-19/France/ARA-SC315/2020\|EPI_ISL_649949\|2020- | .................... | .................... | ..................... | .................... | .................... | .................... |
| S | 785 hCoV-19/France/ARA-SC316/2020\|EPI_ISL_649950\|2020- | .................... | .................... | ..................... | .................... | .................... | .................... |
| S | 786 hCoV-19/France/ARA-SC317/2020\|EPI_ISL_649951\|2020- | .................... | .................... | ..................... | .................... | .................... | .................... |
| S | 787 hCoV-19/France/ARA-SC318/2020\|EPI_ISL_649952\|2020- | .................... | .................... | ..................... | .................... | .................... | .................... |
| S | 788 hCoV-19/France/ARA-SC320/2020\|EPI_ISL_649953\|2020- | .................... | .................... | ..................... | .................... | .................... | .................... |
| S | 789 hCoV-19/France/NAQ-SC198/2020\|EPI_ISL_649954\|2020- | .................... | .................... | ..................... | .................... | .................... | .................... |
| S | 790 hCoV-19/France/NAQ-SC199/2020\|EPI_ISL_649955\|2020- | .................... | .................... | ..................... | .................... | .................... | .................... |
| S | 791 hCoV-19/France/NAQ-SC200/2020\|EPI_ISL_649956\|2020- | .................... | .................... | ..................... | .................... | .................... | .................... |
| S | 792 hCoV-19/France/NAQ-SC202/2020\|EPI_ISL_649957\|2020- | .................... | .................... | ..................... | .................... | .................... | .................... |
| S | 793 hCoV-19/France/NAQ-SC204/2020\|EPI_ISL_649958\|2020- | .................... | .................... | ..................... | .................... | .................... | .................... |
| S | 794 hCoV-19/France/NAQ-SC205/2020\|EPI_ISL_649959\|2020- | .................... | .................... | ..................... | .................... | .................... | .................... |
| S | 795 hCoV-19/France/NAQ-SC206/2020\|EPI_ISL_649960\|2020- | .................... | .................... | ..................... | .................... | .................... | .................... |
| S | 796 hCoV-19/France/NAQ-SC207/2020\|EPI_ISL_649961\|2020- | .................... | .................... | ..................... | .................... | .................... | .................... |
| S | 797 hCoV-19/France/NAQ-SC210/2020\|EPI_ISL_649962\|2020- | .................... | .................... | ..................... | .................... | .................... | .................... |
| S | 798 hCoV-19/France/NAQ-SC211/2020\|EPI_ISL_649963\|2020- | .................... | .................... | ..................... | .................... | .................... | .................... |
| S | 799 hCoV-19/France/NAQ-SC212/2020\|EPI_ISL_649964\|2020- | .................... | .................... | ..................... | .................... | .................... | .................... |
| S | 800 hCoV-19/France/NAQ-SC213/2020\|EPI_ISL_649965\|2020- | .................... | .................... | ..................... | .................... | .................... | .................... |
| S | 801 hCoV-19/France/NAQ-SC214/2020\|EPI_ISL_649966\|2020- | .................... | .................... | ..................... | .................... | .................... | .................... |
| S | 802 hCoV-19/France/NAQ-SC215/2020\|EPI_ISL_649967\|2020- | .................... | .................... | ..................... | .................... | .................... | .................... |
| S | 803 hCoV-19/France/NAQ-SC217/2020\|EPI_ISL_649968\|2020- | .................... | .................... | ..................... | .................... | .................... | .................... |
| S | 804 hCoV-19/France/NAQ-SC218/2020\|EPI_ISL_649969\|2020- | .................... | .................... | ..................... | .................... | .................... | .................... |
| S | 805 hCoV-19/USA/FL-BPHL-1947/2020\|EPI_ISL_653100\|2020- | .................... | .................... | ..................... | .................... | .................... | .................... |
| S | 806 hCoV-19/USA/FL-BPHL-1948/2020\|EPI_ISL_653101\|2020- | .................... | .................... | ..................... | .................... | .................... | .................... |
| S | 807 hCoV-19/USA/FL-BPHL-1949/2020\|EPI_ISL_653102\|2020- | .................... | .................... | ..................... | .................... | .................... | .................... |
| S | 808 hCoV-19/USA/FL-BPHL-1950/2020\|EPI_ISL_653103\|2020- | .................... | .................... | ..................... | .................... | .................... | .................... |
| S | 809 hCoV-19/USA/FL-BPHL-1951/2020\|EPI_ISL_653104\|2020- | .................... | .................... | ..................... | .................... | .................... | .................... |
| S | 810 hCoV-19/USA/FL-BPHL-1952/2020\|EPI_ISL_653105\|2020- | .................... | .................... | ..................... | .................... | .................... | .................... |
| S | 811 hCoV-19/USA/FL-BPHL-1953/2020\|EPI_ISL_653106\|2020- | .................... | .................... | ..................... | .................... | .................... | .................... |
| S | 812 hCoV-19/USA/FL-BPHL-1954/2020\|EPI_ISL_653107\|2020- | .................... | .................... | ..................... | .................... | .................... | .................... |
| S | 814 hCoV-19/USA/FL-BPHL-1956/2020\|EPI_ISL_653109\|2020- | .................... | .................... | ..................... | .................... | .................... | .................... |
| S | 815 hCoV-19/USA/FL-BPHL-1957/2020\|EPI_ISL_653110\|2020- | .................... | .................... | ..................... | .................... | .................... | .................... |
| S | 816 hCoV-19/USA/FL-BPHL-1958/2020\|EPI_ISL_653111\|2020- | .................... | .................... | ..................... | .................... | .................... | .................... |
| S | 817 hCoV-19/USA/FL-BPHL-1959/2020\|EPI_ISL_653112\|2020- | .................... | .................... | ..................... | .................... | .................... | .................... |
| S | 818 hCoV-19/USA/FL-BPHL-1960/2020\|EPI_ISL_653113\|2020- | .................... | .................... | ..................... | .................... | .................... | .................... |
| S | 819 hCoV-19/USA/FL-BPHL-1961/2020\|EPI_ISL_653114\|2020- | .................... | .................... | ..................... | .................... | .................... | .................... |
| S | 820 hCoV-19/USA/FL-BPHL-1962/2020\|EPI_ISL_653115\|2020- | .................... | .................... | ..................... | .................... | .................... | .................... |
| S | 821 hCoV-19/USA/FL-BPHL-1964/2020\|EPI_ISL_653117\|2020- | .................... | .................... | ..................... | .................... | .................... | .................... |
| S | 822 hCoV-19/USA/FL-BPHL-1965/2020\|EPI_ISL_653118\|2020- | .................... | .................... | ..................... | .................... | .................... | .................... |
| S | 823 hCoV-19/USA/FL-BPHL-1966/2020\|EPI_ISL_653119\|2020- | .................... | .................... | ..................... | .................... | .................... | .................... |
| S | 824 hCoV-19/USA/FL-BPHL-1967/2020\|EPI_ISL_653120\|2020- | .................... | .................... | ..................... | .................... | .................... | .................... |
| S | 825 hCoV-19/USA/FL-BPHL-1969/2020\|EPI_ISL_653122\|2020- | .................... | .................... | ..................... | .................... | .................... | .................... |
| S | 826 hCoV-19/USA/FL-BPHL-1970/2020\|EPI_ISL_653123\|2020- | .................... | .................... | ..................... | .................... | .................... | .................... |
| S | 827 hCoV-19/USA/FL-BPHL-1971/2020\|EPI_ISL_653124\|2020- | .................... | .................... | ..................... | .................... | .................... | .................... |
| S | 828 hCoV-19/USA/FL-BPHL-1973/2020\|EPI_ISL_653126\|2020- | .................... | .................... | ..................... | .................... | .................... | .................... |

| | | | | | | | |
|---|---|---|---|---|---|---|---|
| S | 829 hCoV-19/USA/FL-BPHL-1974/2020\|EPI_ISL_653127\|2020- | ···················· | ···················· | ····················· | ···················· | ···················· | ···················· |
| S | 831 hCoV-19/USA/FL-BPHL-1977/2020\|EPI_ISL_653130\|2020- | ···················· | ···················· | ····················· | ···················· | ···················· | ···················· |
| S | 832 hCoV-19/USA/FL-BPHL-1979/2020\|EPI_ISL_653132\|2020- | ···················· | ···················· | ····················· | ···················· | ···················· | ···················· |
| S | 833 hCoV-19/USA/FL-BPHL-1980/2020\|EPI_ISL_653133\|2020- | ···················· | ···················· | ····················· | ···················· | ···················· | ···················· |
| S | 834 hCoV-19/USA/FL-BPHL-1984/2020\|EPI_ISL_653137\|2020- | ···················· | ···················· | ····················· | ···················· | ···················· | ···················· |
| S | 835 hCoV-19/USA/FL-BPHL-1986/2020\|EPI_ISL_653139\|2020- | ···················· | ···················· | ····················· | ···················· | ···················· | ···················· |
| S | 836 hCoV-19/USA/FL-BPHL-1987/2020\|EPI_ISL_653140\|2020- | ···················· | ···················· | ····················· | ···················· | ···················· | ···················· |
| S | 837 hCoV-19/USA/FL-BPHL-1988/2020\|EPI_ISL_653141\|2020- | ···················· | ···················· | ····················· | ···················· | ···················· | ···················· |
| S | 838 hCoV-19/USA/FL-BPHL-1992/2020\|EPI_ISL_653145\|2020- | ···················· | ···················· | ····················· | ···················· | ···················· | ···················· |
| S | 839 hCoV-19/USA/FL-BPHL-1993/2020\|EPI_ISL_653146\|2020- | ···················· | ···················· | ····················· | ···················· | ···················· | ···················· |
| S | 840 hCoV-19/USA/FL-BPHL-1994/2020\|EPI_ISL_653147\|2020- | ···················· | ···················· | ····················· | ···················· | ···················· | ···················· |
| S | 841 hCoV-19/USA/FL-BPHL-1995/2020\|EPI_ISL_653148\|2020- | ···················· | ···················· | ····················· | ···················· | ···················· | ···················· |
| S | 842 hCoV-19/USA/FL-BPHL-2000/2020\|EPI_ISL_653153\|2020- | ···················· | ···················· | ····················· | ···················· | ···················· | ···················· |
| S | 844 hCoV-19/USA/FL-BPHL-2009/2020\|EPI_ISL_653162\|2020- | ···················· | ···················· | ····················· | ···················· | ···················· | ···················· |
| S | 846 hCoV-19/USA/FL-BPHL-2011/2020\|EPI_ISL_653164\|2020- | ···················· | ···················· | ····················· | ···················· | ···················· | ···················· |
| S | 847 hCoV-19/USA/FL-BPHL-2016/2020\|EPI_ISL_653169\|2020- | ···················· | ···················· | ····················· | ···················· | ···················· | ···················· |
| S | 848 hCoV-19/USA/FL-BPHL-2017/2020\|EPI_ISL_653170\|2020- | ···················· | ···················· | ····················· | ···················· | ···················· | ···················· |
| S | 849 hCoV-19/USA/FL-BPHL-2028/2020\|EPI_ISL_653181\|2020- | ···················· | ···················· | ····················· | ···················· | ···················· | ···················· |
| S | 850 hCoV-19/USA/FL-BPHL-2032/2020\|EPI_ISL_653185\|2020- | ···················· | ···················· | ····················· | ···················· | ···················· | ···················· |
| S | 851 hCoV-19/USA/FL-BPHL-2038/2020\|EPI_ISL_653191\|2020- | ···················· | ···················· | ····················· | ···················· | ···················· | ···················· |
| S | 852 hCoV-19/USA/FL-BPHL-2040/2020\|EPI_ISL_653193\|2020- | ···················· | ···················· | ····················· | ···················· | ···················· | ···················· |
| S | 853 hCoV-19/Spain/MD-IBV-99009675/2020\|EPI_ISL_654400\| | ···················· | ···················· | ····················· | ···················· | ···················· | ···················· |
| S | 854 hCoV-19/Spain/AR-IBV-98008011/2020\|EPI_ISL_654421\| | ···················· | ···················· | ····················· | ···················· | ···················· | ···················· |
| S | 855 hCoV-19/Spain/AR-IBV-98008013/2020\|EPI_ISL_654422\| | ···················· | ···················· | ····················· | ···················· | ···················· | ···················· |
| S | 856 hCoV-19/Spain/AR-IBV-98008014/2020\|EPI_ISL_654423\| | ···················· | ···················· | ····················· | ···················· | ···················· | ···················· |
| S | 857 hCoV-19/Spain/AR-IBV-98008066/2020\|EPI_ISL_654424\| | ···················· | ···················· | ····················· | ···················· | ···················· | ···················· |
| S | 858 hCoV-19/Spain/AR-IBV-98008080/2020\|EPI_ISL_654425\| | ···················· | ···················· | ····················· | ···················· | ···················· | ···················· |
| S | 859 hCoV-19/Spain/AR-IBV-98008122/2020\|EPI_ISL_654426\| | ···················· | ···················· | ····················· | ···················· | ···················· | ···················· |
| S | 860 hCoV-19/Spain/AR-IBV-98008123/2020\|EPI_ISL_654427\| | ···················· | ···················· | ····················· | ···················· | ···················· | ···················· |
| S | 861 hCoV-19/Spain/AR-IBV-98008124/2020\|EPI_ISL_654428\| | ···················· | ···················· | ····················· | ···················· | ···················· | ···················· |
| S | 862 hCoV-19/Spain/AR-IBV-98008136/2020\|EPI_ISL_654429\| | ···················· | ···················· | ····················· | ···················· | ···················· | ···················· |
| S | 863 hCoV-19/Spain/AR-IBV-98008146/2020\|EPI_ISL_654430\| | ···················· | ···················· | ····················· | ···················· | ···················· | ···················· |
| S | 864 hCoV-19/Spain/AR-IBV-98008150/2020\|EPI_ISL_654431\| | ···················· | ···················· | ····················· | ···················· | ···················· | ···················· |
| S | 865 hCoV-19/Spain/AR-IBV-98008158/2020\|EPI_ISL_654432\| | ···················· | ···················· | ····················· | ···················· | ···················· | ···················· |
| S | 866 hCoV-19/Spain/AR-IBV-98008038/2020\|EPI_ISL_654441\| | ···················· | ···················· | ····················· | ···················· | ···················· | ···················· |
| S | 867 hCoV-19/Spain/AR-IBV-98008039/2020\|EPI_ISL_654442\| | ···················· | ···················· | ····················· | ···················· | ···················· | ···················· |
| S | 868 hCoV-19/Spain/AR-IBV-98008041/2020\|EPI_ISL_654443\| | ···················· | ···················· | ····················· | ···················· | ···················· | ···················· |
| S | 869 hCoV-19/Spain/AR-IBV-98008009/2020\|EPI_ISL_654444\| | ···················· | ···················· | ····················· | ···················· | ···················· | ···················· |
| S | 870 hCoV-19/Spain/AR-IBV-98008012/2020\|EPI_ISL_654445\| | ···················· | ···················· | ····················· | ···················· | ···················· | ···················· |
| S | 871 hCoV-19/Spain/AR-IBV-98008021/2020\|EPI_ISL_654446\| | ···················· | ···················· | ····················· | ···················· | ···················· | ···················· |
| S | 872 hCoV-19/Spain/AR-IBV-98008036/2020\|EPI_ISL_654447\| | ···················· | ···················· | ····················· | ···················· | ···················· | ···················· |
| S | 873 hCoV-19/Spain/AR-IBV-98008055/2020\|EPI_ISL_654448\| | ···················· | ···················· | ····················· | ···················· | ···················· | ···················· |
| S | 874 hCoV-19/Spain/AR-IBV-98008056/2020\|EPI_ISL_654449\| | ···················· | ···················· | ····················· | ···················· | ···················· | ···················· |
| S | 875 hCoV-19/Spain/AR-IBV-98008059/2020\|EPI_ISL_654450\| | ···················· | ···················· | ····················· | ···················· | ···················· | ···················· |
| S | 876 hCoV-19/Spain/AR-IBV-98008076/2020\|EPI_ISL_654451\| | ···················· | ···················· | ····················· | ···················· | ···················· | ···················· |
| S | 877 hCoV-19/Spain/AR-IBV-98008077/2020\|EPI_ISL_654452\| | ···················· | ···················· | ····················· | ···················· | ···················· | ···················· |
| S | 878 hCoV-19/Spain/AR-IBV-98008079/2020\|EPI_ISL_654453\| | ···················· | A··················· | ····················· | ···················· | ···················· | ···················· |
| S | 879 hCoV-19/Spain/AR-IBV-98008085/2020\|EPI_ISL_654454\| | ···················· | ···················· | ····················· | ···················· | ···················· | ···················· |
| S | 880 hCoV-19/Spain/AR-IBV-98008093/2020\|EPI_ISL_654455\| | ···················· | ···················· | ····················· | ···················· | ···················· | ···················· |
| S | 881 hCoV-19/Spain/AR-IBV-98008094/2020\|EPI_ISL_654456\| | ···················· | ···················· | ····················· | ···················· | ···················· | ···················· |
| S | 882 hCoV-19/Spain/AR-IBV-98008119/2020\|EPI_ISL_654457\| | ···················· | ···················· | ····················· | ···················· | ···················· | ···················· |
| S | 884 hCoV-19/Spain/AR-IBV-98008126/2020\|EPI_ISL_654459\| | ···················· | ···················· | ····················· | ···················· | ···················· | ···················· |
| S | 885 hCoV-19/Spain/AR-IBV-98008128/2020\|EPI_ISL_654460\| | ···················· | ···················· | ····················· | ···················· | ···················· | ···················· |
| S | 886 hCoV-19/Spain/AR-IBV-98008132/2020\|EPI_ISL_654461\| | ···················· | ···················· | ····················· | ···················· | ······T············· | ···················· |
| S | 887 hCoV-19/Spain/AR-IBV-98008134/2020\|EPI_ISL_654462\| | ···················· | ···················· | ····················· | ···················· | ···················· | ···················· |
| S | 888 hCoV-19/Spain/AR-IBV-98008140/2020\|EPI_ISL_654463\| | ···················· | ···················· | ····················· | ···················· | ······T············· | ···················· |
| S | 889 hCoV-19/Spain/AR-IBV-98008141/2020\|EPI_ISL_654464\| | ···················· | ···················· | ····················· | ···················· | ···················· | ···················· |
| S | 890 hCoV-19/Spain/AR-IBV-98008152/2020\|EPI_ISL_654465\| | ···················· | ···················· | ····················· | ···················· | ···················· | ···················· |
| S | 891 hCoV-19/Spain/AR-IBV-98008164/2020\|EPI_ISL_654466\| | ···················· | ···················· | ····················· | ···················· | ···················· | ···················· |
| S | 893 hCoV-19/Spain/VC-IBV-98004324/2020\|EPI_ISL_654469\| | ···················· | ···················· | ····················· | ···················· | ···················· | ···················· |
| S | 894 hCoV-19/Spain/VC-IBV-98004325/2020\|EPI_ISL_654470\| | ···················· | ···················· | ····················· | ···················· | ···················· | ···················· |
| S | 895 hCoV-19/Spain/VC-IBV-98004340/2020\|EPI_ISL_654477\| | ···················· | ···················· | ····················· | ···················· | ···················· | ···················· |
| S | 896 hCoV-19/Spain/VC-IBV-98004342/2020\|EPI_ISL_654478\| | ···················· | ···················· | ····················· | ···················· | ···················· | ···················· |
| S | 897 hCoV-19/Spain/VC-IBV-98007308/2020\|EPI_ISL_654486\| | ···················· | ···················· | ····················· | ···················· | ···················· | ···················· |
| S | 898 hCoV-19/Spain/VC-IBV-98007320/2020\|EPI_ISL_654489\| | ···················· | ···················· | ····················· | ···················· | ···················· | ···················· |
| S | 899 hCoV-19/Spain/VC-IBV-98008001/2020\|EPI_ISL_654497\| | ···················· | ···················· | ····················· | ···················· | ···················· | ···················· |
| S | 900 hCoV-19/Sweden/20-53090/2020\|EPI_ISL_654498\|2020-0 | ···················· | ···················· | ····················· | ···················· | ···················· | ···················· |
| S | 901 hCoV-19/Sweden/20-09856/2020\|EPI_ISL_654499\|2020-1 | ···················· | ···················· | ····················· | ···················· | ···················· | ···················· |
| S | 902 hCoV-19/England/ALDP-B5C86C/2020\|EPI_ISL_655724\|20 | ···················· | ···················· | ····················· | ···················· | ···················· | ···················· |
| S | 903 hCoV-19/England/ALDP-B5CC51/2020\|EPI_ISL_655725\|20 | ···················· | ···················· | ····················· | ···················· | ···················· | ···················· |
| S | 904 hCoV-19/England/ALDP-B5CFA3/2020\|EPI_ISL_655726\|20 | ···················· | ···················· | ····················· | ···················· | ···················· | ···················· |
| S | 905 hCoV-19/England/QEUH-B65F7A/2020\|EPI_ISL_655727\|20 | ···················· | ···················· | ····················· | ···················· | ···················· | ···················· |
| S | 906 hCoV-19/England/ALDP-B5C28C/2020\|EPI_ISL_655728\|20 | ···················· | ···················· | ····················· | ···················· | ···················· | ···················· |
| S | 907 hCoV-19/England/ALDP-B5C0FB/2020\|EPI_ISL_655729\|20 | ···················· | ·A·················· | ····················· | ···················· | ···················· | ···A················ |
| S | 908 hCoV-19/England/ALDP-B5CC24/2020\|EPI_ISL_655730\|20 | ···················· | ···················· | ····················· | ···················· | ···················· | ···················· |
| S | 909 hCoV-19/England/ALDP-B5BCF8/2020\|EPI_ISL_655731\|20 | ···················· | ···················· | ····················· | ···················· | ···················· | ···················· |
| S | 910 hCoV-19/England/ALDP-B5C7F6/2020\|EPI_ISL_655733\|20 | ···················· | ···················· | ····················· | ···················· | ···················· | ···················· |
| S | 911 hCoV-19/England/ALDP-B5C36B/2020\|EPI_ISL_655734\|20 | ···················· | ···················· | ····················· | ···················· | ···················· | ···················· |
| S | 912 hCoV-19/England/QEUH-B6C7D5/2020\|EPI_ISL_655735\|20 | ···················· | ···················· | ····················· | ···················· | ···················· | ···················· |
| S | 913 hCoV-19/England/ALDP-B5C5ED/2020\|EPI_ISL_655736\|20 | ···················· | ···················· | ····················· | ···················· | ···················· | ···················· |

| | | | | | | | |
|---|---|---|---|---|---|---|---|
| S | 914 hCoV-19/England/QEUH-B66049/2020\|EPI_ISL_655737\|20 | ···················· | ···················· | ····················· | ···················· | ···················· | ···················· |
| S | 915 hCoV-19/England/ALDP-B5C0CE/2020\|EPI_ISL_655738\|20 | ···················· | ·A·················· | ····················· | ···················· | ···················· | ···A················ |
| S | 916 hCoV-19/England/QEUH-B67840/2020\|EPI_ISL_655739\|20 | ···················· | ···················· | ····················· | ···················· | ···················· | ···················· |
| S | 917 hCoV-19/England/ALDP-B5CEB5/2020\|EPI_ISL_655741\|20 | ···················· | ···················· | ····················· | ···················· | ···················· | ···················· |
| S | 918 hCoV-19/England/ALDP-B5CCCA/2020\|EPI_ISL_655742\|20 | ···················· | ···················· | ····················· | ···················· | ···················· | ···················· |
| S | 919 hCoV-19/England/ALDP-B5C389/2020\|EPI_ISL_655743\|20 | ···················· | ···················· | ····················· | ···················· | ···················· | ···················· |
| S | 920 hCoV-19/England/QEUH-B6C77B/2020\|EPI_ISL_655744\|20 | ···················· | ···················· | ····················· | ···················· | ···················· | ···················· |
| S | 921 hCoV-19/England/QEUH-B65F01/2020\|EPI_ISL_655745\|20 | ···················· | ···················· | ····················· | ···················· | ···················· | ···················· |
| S | 922 hCoV-19/England/ALDP-B5C4FF/2020\|EPI_ISL_655746\|20 | ···················· | ···················· | ····················· | ···················· | ···················· | ···················· |
| S | 923 hCoV-19/England/ALDP-B5C811/2020\|EPI_ISL_655747\|20 | ···················· | ···················· | ····················· | ···················· | ···················· | ···················· |
| S | 924 hCoV-19/England/ALDP-B5CA66/2020\|EPI_ISL_655748\|20 | ···················· | ···················· | ····················· | ···················· | ···················· | ···················· |
| S | 925 hCoV-19/England/ALDP-B5C5B0/2020\|EPI_ISL_655749\|20 | ···················· | ···················· | ····················· | ···················· | ···················· | ···················· |
| S | 926 hCoV-19/England/ALDP-B5C073/2020\|EPI_ISL_655750\|20 | ···················· | ·A·················· | ····················· | ···················· | ···················· | ···A················ |
| S | 927 hCoV-19/England/ALDP-B5BFE0/2020\|EPI_ISL_655751\|20 | ···················· | ···················· | ····················· | ···················· | ···················· | ···················· |
| S | 928 hCoV-19/England/ALDP-B5C574/2020\|EPI_ISL_655752\|20 | ···················· | ···················· | ····················· | ···················· | ···················· | ···················· |
| S | 929 hCoV-19/England/ALDP-B5CE1F/2020\|EPI_ISL_655753\|20 | ···················· | ···················· | ····················· | ···················· | ···················· | ···················· |
| S | 930 hCoV-19/England/ALDP-B5BF1D/2020\|EPI_ISL_655754\|20 | ···················· | ···················· | ····················· | ···················· | ···················· | ···················· |
| S | 931 hCoV-19/England/ALDP-B5C1CB/2020\|EPI_ISL_655755\|20 | ···················· | ···················· | ····················· | ···················· | ···················· | ···················· |
| S | 932 hCoV-19/England/QEUH-B65E22/2020\|EPI_ISL_655756\|20 | ···················· | ···················· | ····················· | ···················· | ···················· | ···················· |
| S | 933 hCoV-19/England/ALDP-B5BE6B/2020\|EPI_ISL_655757\|20 | ···················· | ···················· | ····················· | ···················· | ···················· | ···················· |
| S | 934 hCoV-19/England/ALDP-B5CBEB/2020\|EPI_ISL_655758\|20 | ···················· | ···················· | ····················· | ···················· | ···················· | ···················· |
| S | 935 hCoV-19/England/ALDP-B5C301/2020\|EPI_ISL_655759\|20 | ···················· | ···················· | ····················· | ···················· | ···················· | ···················· |
| S | 936 hCoV-19/England/ALDP-B5C77E/2020\|EPI_ISL_655760\|20 | ···················· | ···················· | ····················· | ···················· | ······T············· | ···················· |
| S | 937 hCoV-19/England/ALDP-B5CDD6/2020\|EPI_ISL_655761\|20 | ···················· | ·A·················· | ····················· | ···················· | ···················· | ···A················ |
| S | 938 hCoV-19/England/ALDP-B5C1E9/2020\|EPI_ISL_655763\|20 | ···················· | ···················· | ····················· | ···················· | ···················· | ···················· |
| S | 939 hCoV-19/England/ALDP-B5C94B/2020\|EPI_ISL_655764\|20 | ···················· | ···················· | ····················· | ···················· | ···················· | ···················· |
| S | 940 hCoV-19/England/ALDP-B5C9E1/2020\|EPI_ISL_655767\|20 | ···················· | ···················· | ····················· | ···················· | ······T············· | ···················· |
| S | 941 hCoV-19/England/QEUH-B6C6C9/2020\|EPI_ISL_655768\|20 | ···················· | ···················· | ····················· | ···················· | ···················· | ···················· |
| S | 942 hCoV-19/England/QEUH-B6600D/2020\|EPI_ISL_655769\|20 | ···················· | ···················· | ····················· | ···················· | ···················· | ···················· |
| S | 943 hCoV-19/England/ALDP-B5C00A/2020\|EPI_ISL_655771\|20 | ···················· | ···················· | ····················· | ···················· | ···················· | ···················· |
| S | 944 hCoV-19/England/QEUH-B678AA/2020\|EPI_ISL_655773\|20 | ···················· | ···················· | ····················· | ···················· | ···················· | ···················· |
| S | 945 hCoV-19/England/ALDP-B5C3D4/2020\|EPI_ISL_655774\|20 | ···················· | ···················· | ····················· | ···················· | ···················· | ···················· |
| S | 946 hCoV-19/England/ALDP-B5C6EA/2020\|EPI_ISL_655775\|20 | ···················· | ···················· | ····················· | ···················· | ···················· | ···················· |
| S | 947 hCoV-19/England/ALDP-B5C3F2/2020\|EPI_ISL_655776\|20 | ···················· | ···················· | ····················· | ···················· | ···················· | ···················· |
| S | 948 hCoV-19/England/ALDP-B5CC15/2020\|EPI_ISL_655778\|20 | ···················· | ···················· | ····················· | ···················· | ···················· | ···················· |
| S | 949 hCoV-19/England/QEUH-B6C81E/2020\|EPI_ISL_655779\|20 | ···················· | ···················· | ····················· | ···················· | ···················· | ···················· |
| S | 950 hCoV-19/England/QEUH-B6C7C6/2020\|EPI_ISL_655780\|20 | ···················· | ···················· | ····················· | ···················· | ···················· | ···················· |
| S | 951 hCoV-19/England/ALDP-B5C8A8/2020\|EPI_ISL_655781\|20 | ···················· | ···················· | ····················· | ···················· | ···················· | ···················· |
| S | 952 hCoV-19/England/QEUH-B65EF5/2020\|EPI_ISL_655783\|20 | ···················· | ···················· | ····················· | ···················· | ···················· | ···················· |
| S | 953 hCoV-19/England/ALDP-B5C6F9/2020\|EPI_ISL_655784\|20 | ···················· | ···················· | ····················· | ···················· | ···················· | ···················· |
| S | 954 hCoV-19/England/ALDP-B5C547/2020\|EPI_ISL_655785\|20 | ···················· | ···················· | ····················· | ···················· | ···················· | ···················· |
| S | 955 hCoV-19/England/ALDP-B5C653/2020\|EPI_ISL_655786\|20 | ···················· | ···················· | ····················· | ···················· | ···················· | ···················· |
| S | 956 hCoV-19/England/ALDP-B5C538/2020\|EPI_ISL_655787\|20 | ···················· | ···················· | ····················· | ···················· | ···················· | ···················· |
| S | 957 hCoV-19/England/QEUH-B65EC8/2020\|EPI_ISL_655788\|20 | ···················· | ···················· | ····················· | ···················· | ···················· | ···················· |
| S | 958 hCoV-19/England/ALDP-B5C3B6/2020\|EPI_ISL_655789\|20 | ···················· | ···················· | ····················· | ···················· | ···················· | ···················· |
| S | 959 hCoV-19/England/ALDP-B5BFFF/2020\|EPI_ISL_655790\|20 | ···················· | ···················· | ····················· | ···················· | ···················· | ···················· |
| S | 960 hCoV-19/England/ALDP-B5BF59/2020\|EPI_ISL_655791\|20 | ···················· | ···················· | ····················· | ···················· | ···················· | ···················· |
| S | 961 hCoV-19/England/ALDP-B5C987/2020\|EPI_ISL_655792\|20 | ···················· | ···················· | ····················· | ···················· | ···················· | ···················· |
| S | 962 hCoV-19/England/ALDP-B5BCDA/2020\|EPI_ISL_655793\|20 | ···················· | ···················· | ····················· | ···················· | ······T············· | ···················· |
| S | 963 hCoV-19/England/ALDP-B5C37A/2020\|EPI_ISL_655794\|20 | ···················· | ···················· | ····················· | ···················· | ···················· | ···················· |
| S | 964 hCoV-19/England/ALDP-B5C90F/2020\|EPI_ISL_655795\|20 | ···················· | ···················· | ····················· | ···················· | ···················· | ···················· |
| S | 965 hCoV-19/England/ALDP-B5CF49/2020\|EPI_ISL_655796\|20 | ···················· | ·········T·········· | ····················· | ···················· | ···················· | ···················· |
| S | 966 hCoV-19/England/ALDP-B5C91E/2020\|EPI_ISL_655797\|20 | ···················· | ···················· | ····················· | ···················· | ···················· | ···················· |
| S | 967 hCoV-19/England/QEUH-B6C82D/2020\|EPI_ISL_655798\|20 | ···················· | ···················· | ····················· | ···················· | ···················· | ···················· |
| S | 968 hCoV-19/England/ALDP-B5C7AB/2020\|EPI_ISL_655799\|20 | ···················· | ···················· | ····················· | ···················· | ···················· | ···················· |
| S | 969 hCoV-19/England/MILK-B54AD7/2020\|EPI_ISL_658058\|20 | ···················· | ···················· | ····················· | ···················· | ···················· | ···················· |
| S | 970 hCoV-19/England/MILK-B54B2F/2020\|EPI_ISL_658059\|20 | ···················· | ···················· | ····················· | ···················· | ···················· | ···A················ |
| S | 971 hCoV-19/England/CAMC-B336DA/2020\|EPI_ISL_659211\|20 | ···················· | ···················· | ····················· | ···················· | ···················· | ···················· |
| S | 972 hCoV-19/England/CAMC-B3379B/2020\|EPI_ISL_659212\|20 | ···················· | ···················· | ····················· | ···················· | ···················· | ···················· |
| S | 973 hCoV-19/England/CAMC-B33573/2020\|EPI_ISL_659213\|20 | ···················· | ···················· | ····················· | ···················· | ···················· | ···················· |
| S | 974 hCoV-19/England/CAMC-B338C5/2020\|EPI_ISL_659214\|20 | ···················· | ···················· | ····················· | ···················· | ···················· | ···················· |
| S | 975 hCoV-19/England/CAMC-B336CB/2020\|EPI_ISL_659215\|20 | ···················· | ···················· | ····················· | ···················· | ···················· | ···················· |
| S | 976 hCoV-19/England/CAMC-B336E9/2020\|EPI_ISL_659216\|20 | ···················· | ···················· | ····················· | ···················· | ···················· | ···A················ |
| S | 977 hCoV-19/England/CAMC-B32240/2020\|EPI_ISL_659217\|20 | ···················· | ···················· | ····················· | ···················· | ···················· | ···················· |
| S | 978 hCoV-19/England/CAMC-B3226E/2020\|EPI_ISL_659218\|20 | ···················· | ···················· | ····················· | ···················· | ···················· | ···················· |
| S | 979 hCoV-19/England/CAMC-B33959/2020\|EPI_ISL_659219\|20 | ···················· | ···················· | ····················· | ···················· | ···················· | ···················· |
| S | 980 hCoV-19/England/CAMC-B32EF1/2020\|EPI_ISL_659220\|20 | ···················· | ···················· | ····················· | ···················· | ···················· | ···················· |
| S | 981 hCoV-19/England/CAMC-B33616/2020\|EPI_ISL_659221\|20 | ···················· | ···················· | ····················· | ···················· | ···················· | ···················· |
| S | 982 hCoV-19/England/CAMC-B338A7/2020\|EPI_ISL_659222\|20 | ···················· | ···················· | ····················· | ···················· | ······T············· | ···················· |
| S | 983 hCoV-19/England/CAMC-B323E3/2020\|EPI_ISL_659223\|20 | ···················· | ···················· | ····················· | ···················· | ···················· | ···················· |
| S | 984 hCoV-19/England/CAMC-B32556/2020\|EPI_ISL_659224\|20 | ···················· | ···················· | ····················· | ···················· | ···················· | ···················· |
| S | 985 hCoV-19/England/CAMC-B331E8/2020\|EPI_ISL_659225\|20 | ···················· | ···················· | ····················· | ···················· | ···················· | ···················· |
| S | 986 hCoV-19/England/CAMC-B321E9/2020\|EPI_ISL_659227\|20 | ···················· | ···················· | ····················· | ···················· | ···················· | ···················· |
| S | 987 hCoV-19/England/CAMC-B33C50/2020\|EPI_ISL_659229\|20 | ···················· | ···················· | ····················· | ···················· | ···················· | ···················· |
| S | 988 hCoV-19/England/CAMC-B3384D/2020\|EPI_ISL_659230\|20 | ···················· | ···················· | ····················· | ···················· | ···················· | ···················· |
| S | 989 hCoV-19/England/CAMC-B33625/2020\|EPI_ISL_659232\|20 | ···················· | ···················· | ····················· | ···················· | ···················· | ···················· |
| S | 990 hCoV-19/England/CAMC-B32565/2020\|EPI_ISL_659234\|20 | ···················· | ···················· | ····················· | ···················· | ···················· | ···················· |
| S | 991 hCoV-19/England/CAMC-B325B0/2020\|EPI_ISL_659237\|20 | ···················· | ···················· | ····················· | ···················· | ···················· | ···················· |
| S | 992 hCoV-19/England/CAMC-B32F67/2020\|EPI_ISL_659242\|20 | ···················· | ···················· | ····················· | ···················· | ···················· | ···················· |
| S | 993 hCoV-19/England/CAMC-B33652/2020\|EPI_ISL_659244\|20 | ···················· | ···················· | ····················· | ···················· | ···················· | ···················· |

| | | | | | | | |
|---|---|---|---|---|---|---|---|
| S | 994 hCoV-19/England/CAMC-B33D3F/2020\|EPI_ISL_659245\|20 | ···················· | ···················· | ····················· | ···················· | ···················· | ···················· |
| S | 995 hCoV-19/England/CAMC-B33EB4/2020\|EPI_ISL_659246\|20 | ···················· | ···················· | ····················· | ···················· | ···················· | ···················· |
| S | 996 hCoV-19/England/CAMC-B334FE/2020\|EPI_ISL_659247\|20 | ···················· | ···················· | ····················· | ···················· | ···················· | ···················· |
| S | 997 hCoV-19/England/CAMC-B33BDB/2020\|EPI_ISL_659248\|20 | ···················· | ···················· | ····················· | ···················· | ···················· | ···················· |
| S | 998 hCoV-19/England/CAMC-B3331F/2020\|EPI_ISL_659389\|20 | ···················· | ···················· | ····················· | ···················· | ···················· | ···················· |
| S | 999 hCoV-19/South Africa/KRISP-K004334/2020\|EPI_ISL_66 | ···················· | ·A·················· | ····················· | ···················· | ···················· | ···················· |
| S | 1000 hCoV-19/South Africa/KRISP-K004335/2020\|EPI_ISL_66 | ···················· | ···················· | ····················· | ···················· | ···················· | ···················· |
| S | 1001 hCoV-19/South Africa/KRISP-K004336/2020\|EPI_ISL_66 | ···················· | ···················· | ····················· | ···················· | ···················· | ···················· |
| S | 1002 hCoV-19/South Africa/KRISP-K004337/2020\|EPI_ISL_66 | ···················· | ·A·················· | ····················· | ···················· | ···················· | ···················· |
| S | 1003 hCoV-19/South Africa/KRISP-K004338/2020\|EPI_ISL_66 | ···················· | ···················· | ····················· | ···················· | ···················· | ···················· |
| S | 1004 hCoV-19/South Africa/KRISP-K004339/2020\|EPI_ISL_66 | ···················· | ···················· | ····················· | ···················· | ···················· | ···················· |
| S | 1005 hCoV-19/South Africa/KRISP-K004340/2020\|EPI_ISL_66 | ···················· | ···················· | ····················· | ···················· | ···················· | ···················· |
| S | 1006 hCoV-19/South Africa/KRISP-K004341/2020\|EPI_ISL_66 | ···················· | ·A·················· | ····················· | ···················· | ···················· | ···················· |
| S | 1007 hCoV-19/South Africa/KRISP-K004342/2020\|EPI_ISL_66 | ···················· | ···················· | ····················· | ···················· | ···················· | ···················· |
| S | 1008 hCoV-19/South Africa/KRISP-K004344/2020\|EPI_ISL_66 | ···················· | ·A·················· | ····················· | ···················· | ···················· | ···················· |
| S | 1009 hCoV-19/South Africa/KRISP-K004346/2020\|EPI_ISL_66 | ···················· | ···················· | ····················· | ···················· | ···················· | ···················· |
| S | 1010 hCoV-19/South Africa/KRISP-K004347-CS1/2020\|EPI_IS | ···················· | ···················· | ····················· | ···················· | ···················· | ···················· |
| S | 1011 hCoV-19/South Africa/KRISP-K004351-CE1/2020\|EPI_IS | ···················· | ···················· | ····················· | ···················· | ···················· | ···················· |
| S | 1012 hCoV-19/South Africa/KRISP-K004352-CE1/2020\|EPI_IS | ···················· | ···················· | ····················· | ···················· | ···················· | ···················· |
| S | 1014 hCoV-19/South Africa/KRISP-K004365-CE1/2020\|EPI_IS | ···················· | ···················· | ····················· | ···················· | ···················· | ···················· |
| S | 1015 hCoV-19/South Africa/KRISP-K004530-CS1/2020\|EPI_IS | ···················· | ···················· | ····················· | ···················· | ···················· | ···················· |
| S | 1016 hCoV-19/South Africa/KRISP-K004531-CS1/2020\|EPI_IS | ···················· | ···················· | ····················· | ···················· | ···················· | ···················· |
| S | 1017 hCoV-19/South Africa/KRISP-K004532-CS1/2020\|EPI_IS | ···················· | ···················· | ····················· | ···················· | ···················· | ···················· |
| S | 1018 hCoV-19/South Africa/KRISP-K004533-CS1/2020\|EPI_IS | ···················· | ···················· | ····················· | ···················· | ···················· | ···················· |
| S | 1019 hCoV-19/South Africa/KRISP-K004534-CS1/2020\|EPI_IS | ···················· | ···················· | ····················· | ···················· | ···················· | ···················· |
| S | 1020 hCoV-19/France/ARA-SC406/2020\|EPI_ISL_660365\|2020- | ···················· | ·A·················· | ····················· | ···················· | ···················· | ···················· |
| S | 1021 hCoV-19/France/ARA-SC407/2020\|EPI_ISL_660366\|2020- | ···················· | ···················· | ···········T········· | ···················· | ···················· | ···················· |
| S | 1022 hCoV-19/France/ARA-SC408/2020\|EPI_ISL_660367\|2020- | ···················· | ·A·················· | ····················· | ···················· | ···················· | ···················· |
| S | 1023 hCoV-19/France/ARA-SC409/2020\|EPI_ISL_660368\|2020- | ···················· | ···················· | ···········T········· | ···················· | ···················· | ···················· |
| S | 1024 hCoV-19/France/ARA-SC410/2020\|EPI_ISL_660369\|2020- | ···················· | ···················· | ····················· | ···················· | ···················· | ···················· |
| S | 1025 hCoV-19/England/204380861/2020\|EPI_ISL_649849\|2020 | ···················· | ···················· | ····················· | ···················· | ···················· | ···················· |
| S | 1026 hCoV-19/England/204381146/2020\|EPI_ISL_649850\|2020 | ···················· | ···················· | ····················· | ···················· | ···················· | ···················· |
| S | 1027 hCoV-19/England/204381147/2020\|EPI_ISL_649851\|2020 | ···················· | ···················· | ····················· | ···················· | ···················· | ···················· |
| S | 1028 hCoV-19/England/204381148/2020\|EPI_ISL_649852\|2020 | ···················· | ···················· | ····················· | ···················· | ···················· | ···················· |
| S | 1029 hCoV-19/England/204381150/2020\|EPI_ISL_649853\|2020 | ···················· | ···················· | ····················· | ···················· | ···················· | ···················· |
| S | 1030 hCoV-19/England/204381327/2020\|EPI_ISL_649854\|2020 | ···················· | ·A·················· | ····················· | ···················· | ···················· | ···A················ |
| S | 1031 hCoV-19/England/204381329/2020\|EPI_ISL_649855\|2020 | ···················· | ·A·················· | ····················· | ···················· | ···················· | ···A················ |
| S | 1032 hCoV-19/England/204381353/2020\|EPI_ISL_649856\|2020 | ···················· | ···················· | ····················· | ···················· | ···················· | ···················· |
| S | 1033 hCoV-19/England/204381363/2020\|EPI_ISL_649857\|2020 | ···················· | ···················· | ····················· | ···················· | ···················· | ···················· |
| S | 1034 hCoV-19/England/204381368/2020\|EPI_ISL_649858\|2020 | ···················· | ···················· | ····················· | ···················· | ···················· | ···················· |
| S | 1035 hCoV-19/England/204381369/2020\|EPI_ISL_649859\|2020 | ···················· | ···················· | ····················· | ···················· | ···················· | ···················· |
| S | 1036 hCoV-19/England/204381371/2020\|EPI_ISL_649860\|2020 | ···················· | ···················· | ····················· | ···················· | ···················· | ···················· |
| S | 1037 hCoV-19/England/204381377/2020\|EPI_ISL_649861\|2020 | ···················· | ···················· | ····················· | ···················· | ···················· | ···················· |
| S | 1038 hCoV-19/England/204381380/2020\|EPI_ISL_649862\|2020 | ···················· | ···················· | ····················· | ···················· | ···················· | ···················· |
| S | 1039 hCoV-19/England/204381381/2020\|EPI_ISL_649863\|2020 | ···················· | ···················· | ····················· | ···················· | ···················· | ···················· |
| S | 1040 hCoV-19/England/204381382/2020\|EPI_ISL_649864\|2020 | ···················· | ···················· | ····················· | ···················· | ······T············· | ···················· |
| S | 1041 hCoV-19/England/204381398/2020\|EPI_ISL_649865\|2020 | ···················· | ···················· | ····················· | ···················· | ···················· | ···················· |
| S | 1042 hCoV-19/England/204381401/2020\|EPI_ISL_649866\|2020 | ···················· | ···················· | ····················· | ···················· | ···················· | ···················· |
| S | 1043 hCoV-19/England/204390680/2020\|EPI_ISL_649867\|2020 | ···················· | ···················· | ····················· | ···················· | ···················· | ···················· |
| S | 1044 hCoV-19/England/204390683/2020\|EPI_ISL_649869\|2020 | ···················· | ···················· | ····················· | ···················· | ···················· | ···················· |
| S | 1045 hCoV-19/England/204390701/2020\|EPI_ISL_649871\|2020 | ···················· | ···················· | ····················· | ···················· | ···················· | ···················· |
| S | 1046 hCoV-19/England/204390703/2020\|EPI_ISL_649872\|2020 | ···················· | ···················· | ····················· | ···················· | ···················· | ···················· |
| S | 1047 hCoV-19/England/204390706/2020\|EPI_ISL_649873\|2020 | ···················· | ···················· | ····················· | ···················· | ···················· | ···················· |
| S | 1048 hCoV-19/England/204390715/2020\|EPI_ISL_649874\|2020 | ···················· | ···················· | ····················· | ···················· | ···················· | ···················· |
| S | 1049 hCoV-19/England/204390996/2020\|EPI_ISL_649875\|2020 | ···················· | ···················· | ····················· | ···················· | ···················· | ···················· |
| S | 1050 hCoV-19/England/204390997/2020\|EPI_ISL_649876\|2020 | ···················· | ···················· | ····················· | ···················· | ···················· | ···················· |
| S | 1051 hCoV-19/England/204391000/2020\|EPI_ISL_649877\|2020 | ···················· | ···················· | ····················· | ···················· | ···················· | ···················· |
| S | 1052 hCoV-19/England/204391017/2020\|EPI_ISL_649878\|2020 | ···················· | ·A·················· | ····················· | ···················· | ···················· | ···A················ |
| S | 1053 hCoV-19/England/204391027/2020\|EPI_ISL_649879\|2020 | ···················· | ···················· | ····················· | ···················· | ···················· | ···················· |
| S | 1054 hCoV-19/England/204391040/2020\|EPI_ISL_649880\|2020 | ···················· | ···················· | ····················· | ···················· | ···················· | ···················· |
| S | 1055 hCoV-19/England/204393754/2020\|EPI_ISL_649881\|2020 | ···················· | ···················· | ····················· | ···················· | ······T············· | ···················· |
| S | 1056 hCoV-19/England/204393758/2020\|EPI_ISL_649883\|2020 | ···················· | ···················· | ····················· | ···················· | ···················· | ···················· |
| S | 1057 hCoV-19/England/204393759/2020\|EPI_ISL_649884\|2020 | ···················· | ···················· | ····················· | ···················· | ······T············· | ···················· |
| S | 1058 hCoV-19/England/204393809/2020\|EPI_ISL_649885\|2020 | ···················· | ···················· | ····················· | ···················· | ···················· | ···················· |
| S | 1059 hCoV-19/England/204393818/2020\|EPI_ISL_649886\|2020 | ···················· | ···················· | ····················· | ···················· | ···················· | ···················· |
| S | 1060 hCoV-19/England/204393937/2020\|EPI_ISL_649887\|2020 | ···················· | ·A·················· | ····················· | ···················· | ···················· | ···A················ |
| S | 1061 hCoV-19/England/204393980/2020\|EPI_ISL_649888\|2020 | ···················· | ·A·················· | ····················· | ···················· | ···················· | ···A················ |
| S | 1062 hCoV-19/England/204700085/2020\|EPI_ISL_649889\|2020 | ···················· | ···················· | ····················· | ···················· | ···················· | ···················· |
| S | 1063 hCoV-19/England/QEUH-AAEDA9/2020\|EPI_ISL_653006\|20 | ···················· | ·A·················· | ····················· | ···················· | ···················· | ···················· |
| S | 1064 hCoV-19/England/QEUH-AAEDD6/2020\|EPI_ISL_653007\|20 | ···················· | ···················· | ····················· | ···················· | ···················· | ···················· |
| S | 1065 hCoV-19/England/QEUH-AAEDE5/2020\|EPI_ISL_653008\|20 | ···················· | ···················· | ····················· | ···················· | ···················· | ···················· |
| S | 1066 hCoV-19/England/QEUH-AAEDF4/2020\|EPI_ISL_653009\|20 | ···················· | ···················· | ····················· | ···················· | ···················· | ···················· |
| S | 1067 hCoV-19/England/QEUH-AAEFD0/2020\|EPI_ISL_653010\|20 | ···················· | ···················· | ····················· | ···················· | ···················· | ···················· |
| S | 1068 hCoV-19/England/QEUH-AAEFFE/2020\|EPI_ISL_653011\|20 | ···················· | ···················· | ····················· | ···················· | ···················· | ···················· |
| S | 1069 hCoV-19/England/QEUH-AAF045/2020\|EPI_ISL_653012\|20 | ···················· | ···················· | ····················· | ···················· | ····T··············· | ···················· |
| S | 1070 hCoV-19/England/QEUH-AAF063/2020\|EPI_ISL_653013\|20 | ···················· | ···················· | ····················· | ···················· | ···················· | ···················· |
| S | 1071 hCoV-19/Scotland/QEUH-AAF133/2020\|EPI_ISL_653014\|2 | ···················· | ···················· | ····················· | ···················· | ···················· | ···················· |
| S | 1072 hCoV-19/Scotland/QEUH-AAF142/2020\|EPI_ISL_653015\|2 | ···················· | ···················· | ····················· | ···················· | ···················· | ···················· |
| S | 1073 hCoV-19/Scotland/QEUH-AAF18E/2020\|EPI_ISL_653016\|2 | ···················· | ···················· | ····················· | ···················· | ···················· | ···················· |
| S | 1074 hCoV-19/Scotland/QEUH-AAF1E8/2020\|EPI_ISL_653017\|2 | ···················· | ···················· | ····················· | ···················· | ···················· | ···················· |

| | | | | | | | | |
|---|---|---|---|---|---|---|---|---|
| S | 1075 | hCoV-19/Scotland/QEUH-AAF397/2020\|EPI_ISL_653018\|2 | ···················· | ···················· | ···················· | ···················· | ···················· | ···················· |
| S | 1076 | hCoV-19/Scotland/QEUH-AAF3B5/2020\|EPI_ISL_653019\|2 | ···················· | ···················· | ···················· | ···················· | ···················· | ···················· |
| S | 1077 | hCoV-19/Scotland/QEUH-AAF3C4/2020\|EPI_ISL_653020\|2 | ···················· | ···················· | ···················· | ···················· | ···················· | ···················· |
| S | 1078 | hCoV-19/Scotland/QEUH-AAF3F1/2020\|EPI_ISL_653021\|2 | ···················· | ···················· | ···················· | ···················· | ···················· | ···················· |
| S | 1079 | hCoV-19/England/QEUH-AAF42B/2020\|EPI_ISL_653022\|20 | ···················· | ···················· | ···················· | ···················· | ···················· | ···················· |
| S | 1080 | hCoV-19/England/QEUH-AAF43A/2020\|EPI_ISL_653023\|20 | ···················· | ···················· | ···················· | ···················· | ···················· | ···················· |
| S | 1081 | hCoV-19/Scotland/QEUH-AAF449/2020\|EPI_ISL_653024\|2 | ···················· | ···················· | ···················· | ···················· | ···················· | ···················· |
| S | 1082 | hCoV-19/England/QEUH-AAF485/2020\|EPI_ISL_653025\|20 | ···················· | ···················· | ···················· | ···················· | ···················· | ···················· |
| S | 1083 | hCoV-19/England/QEUH-AAF4B2/2020\|EPI_ISL_653026\|20 | ···················· | ···················· | ···················· | ···················· | ···················· | ···················· |
| S | 1084 | hCoV-19/England/QEUH-AF54BD/2020\|EPI_ISL_653027\|20 | ···················· | ·A·················· | ···················· | ···················· | ···················· | ···················· |
| S | 1085 | hCoV-19/Spain/MD-IBV-99008777/2020\|EPI_ISL_654320\| | ···················· | ·····A·············· | ···················· | ···················· | ···················· | ···················· |
| S | 1086 | hCoV-19/Spain/MD-IBV-99008778/2020\|EPI_ISL_654321\| | ···················· | ···················· | ···················· | ···················· | ···················· | ···················· |
| S | 1088 | hCoV-19/Spain/MD-IBV-99008781/2020\|EPI_ISL_654324\| | ···················· | ···················· | ···················· | ···················· | ···················· | ···················· |
| S | 1089 | hCoV-19/Spain/MD-IBV-99008782/2020\|EPI_ISL_654325\| | ···················· | ···················· | ···················· | ···················· | ···················· | ···················· |
| S | 1090 | hCoV-19/Spain/MD-IBV-99009625/2020\|EPI_ISL_654337\| | ···················· | ···················· | ···················· | ···················· | ···················· | ···················· |
| S | 1095 | hCoV-19/Spain/MD-IBV-99009590/2020\|EPI_ISL_654342\| | ···················· | ···················· | ···················· | ···················· | ···················· | ···················· |
| S | 1096 | hCoV-19/Spain/MD-IBV-99009591/2020\|EPI_ISL_654343\| | ···················· | ···················· | ···················· | ···················· | ···················· | ···················· |
| S | 1098 | hCoV-19/Spain/MD-IBV-99009593/2020\|EPI_ISL_654345\| | ···················· | ···················· | ···················· | ···················· | ···················· | ···················· |
| S | 1099 | hCoV-19/Spain/MD-IBV-99009594/2020\|EPI_ISL_654346\| | ···················· | ···················· | ···················· | ···················· | ···················· | ···················· |
| S | 1100 | hCoV-19/Spain/MD-IBV-99009595/2020\|EPI_ISL_654347\| | ···················· | ···················· | ···················· | ···················· | ···················· | ···················· |
| S | 1107 | hCoV-19/Spain/MD-IBV-99009605/2020\|EPI_ISL_654354\| | ······C············· | ···················· | ···················· | ···················· | ···················· | ···················· |
| S | 1112 | hCoV-19/England/ALDP-B5CE6A/2020\|EPI_ISL_655650\|20 | ···················· | ···················· | ···················· | ···················· | ···················· | ···················· |
| S | 1113 | hCoV-19/England/QEUH-B65E7D/2020\|EPI_ISL_655651\|20 | ···················· | ·A·················· | ···················· | ···················· | ···················· | ···················· |
| S | 1114 | hCoV-19/England/ALDP-B5BF0E/2020\|EPI_ISL_655652\|20 | ···················· | ···················· | ···················· | ···················· | ···················· | ···················· |
| S | 1115 | hCoV-19/England/ALDP-B5C9D2/2020\|EPI_ISL_655654\|20 | ···················· | ···················· | ···················· | ···················· | ···················· | ···················· |
| S | 1116 | hCoV-19/England/ALDP-B5C495/2020\|EPI_ISL_655655\|20 | ···················· | ···················· | ···················· | ···················· | ···················· | ···················· |
| S | 1117 | hCoV-19/England/ALDP-B5CF85/2020\|EPI_ISL_655656\|20 | ···················· | ···················· | ···················· | ···················· | ···················· | ···················· |
| S | 1118 | hCoV-19/England/ALDP-B5CE5B/2020\|EPI_ISL_655668\|20 | ···················· | ···················· | ···················· | ···················· | ···················· | ···················· |
| S | 1119 | hCoV-19/England/ALDP-B5CB36/2020\|EPI_ISL_655670\|20 | ···················· | ···················· | ···················· | ···················· | ···················· | ···················· |
| S | 1120 | hCoV-19/England/ALDP-B5C5DE/2020\|EPI_ISL_655671\|20 | ···················· | ···················· | ···················· | ···················· | ···················· | ···················· |
| S | 1121 | hCoV-19/England/QEUH-B6C6F6/2020\|EPI_ISL_655672\|20 | ···················· | ···················· | ···················· | ···················· | ···················· | ···················· |
| S | 1122 | hCoV-19/England/ALDP-B5CB09/2020\|EPI_ISL_655673\|20 | ···················· | ···················· | ···················· | ···················· | ···················· | ···················· |
| S | 1123 | hCoV-19/England/ALDP-B5C046/2020\|EPI_ISL_655674\|20 | ···················· | ···················· | ···················· | ···················· | ···················· | ···················· |
| S | 1124 | hCoV-19/England/ALDP-B5CF94/2020\|EPI_ISL_655675\|20 | ···················· | ···················· | ···················· | ···················· | ···················· | ···················· |
| S | 1125 | hCoV-19/England/QEUH-B6C711/2020\|EPI_ISL_655677\|20 | ···················· | ···················· | ···················· | ···················· | ···················· | ···················· |
| S | 1126 | hCoV-19/England/ALDP-B5C6CC/2020\|EPI_ISL_655678\|20 | ···················· | ···················· | ···················· | ···················· | ···················· | ···················· |
| S | 1127 | hCoV-19/England/ALDP-B5C680/2020\|EPI_ISL_655679\|20 | ···················· | ···················· | ···················· | ···················· | ·····T·············· | ···················· |
| S | 1128 | hCoV-19/England/ALDP-B5CE4C/2020\|EPI_ISL_655680\|20 | ···················· | ···················· | ···················· | ···················· | ···················· | ···················· |
| S | 1129 | hCoV-19/England/ALDP-B5C2D7/2020\|EPI_ISL_655681\|20 | ···················· | ···················· | ···················· | ···················· | ···················· | ···················· |
| S | 1130 | hCoV-19/England/ALDP-B5C79C/2020\|EPI_ISL_655682\|20 | ···················· | ···················· | ···················· | ···················· | ···················· | ···················· |
| S | 1131 | hCoV-19/England/ALDP-B5CF3A/2020\|EPI_ISL_655683\|20 | ···················· | ···················· | ···················· | ···················· | ···················· | ···················· |
| S | 1132 | hCoV-19/England/ALDP-B5C83F/2020\|EPI_ISL_655684\|20 | ···················· | ···················· | ···················· | ···················· | ···················· | ··A················· |
| S | 1133 | hCoV-19/England/ALDP-B5CDB8/2020\|EPI_ISL_655685\|20 | ···················· | ···················· | ···················· | ···················· | ···················· | ···················· |
| S | 1134 | hCoV-19/England/ALDP-B5C7BA/2020\|EPI_ISL_655686\|20 | ···················· | ···················· | ···················· | ···················· | ···················· | ···················· |
| S | 1135 | hCoV-19/England/ALDP-B5C35C/2020\|EPI_ISL_655687\|20 | ···················· | ···················· | ···················· | ···················· | ···················· | ···················· |
| S | 1136 | hCoV-19/England/ALDP-B5C468/2020\|EPI_ISL_655689\|20 | ···················· | ···················· | ···················· | ···················· | ···················· | ···················· |
| S | 1137 | hCoV-19/England/ALDP-B5C626/2020\|EPI_ISL_655700\|20 | ···················· | ···················· | ···················· | ···················· | ···················· | ···················· |
| S | 1138 | hCoV-19/England/ALDP-B5C87B/2020\|EPI_ISL_655701\|20 | ···················· | ···················· | ···················· | ···················· | ···················· | ···················· |
| S | 1139 | hCoV-19/England/ALDP-B5CEE2/2020\|EPI_ISL_655702\|20 | ···················· | ···················· | ···················· | ···················· | ···················· | ···················· |
| S | 1140 | hCoV-19/England/ALDP-B5CC06/2020\|EPI_ISL_655703\|20 | ···················· | ···················· | ···················· | ···················· | ···················· | ···················· |
| S | 1141 | hCoV-19/England/ALDP-B5C125/2020\|EPI_ISL_655704\|20 | ···················· | ·A·················· | ···················· | ···················· | ···················· | ··A················· |
| S | 1142 | hCoV-19/England/ALDP-B5CB81/2020\|EPI_ISL_655705\|20 | ···················· | ···················· | ···················· | ···················· | ···················· | ···················· |
| S | 1143 | hCoV-19/England/ALDP-B5BFC2/2020\|EPI_ISL_655706\|20 | ···················· | ···················· | ···················· | ···················· | ···················· | ···················· |
| S | 1144 | hCoV-19/England/ALDP-B5C40E/2020\|EPI_ISL_655707\|20 | ···················· | ···················· | ···················· | ···················· | ···················· | ···················· |
| S | 1145 | hCoV-19/England/ALDP-B5CB63/2020\|EPI_ISL_655708\|20 | ···················· | ···················· | ···················· | ···················· | ···················· | ···················· |
| S | 1146 | hCoV-19/England/ALDP-B5C1BC/2020\|EPI_ISL_655709\|20 | ···················· | ···················· | ···················· | ···················· | ···················· | ···················· |
| S | 1147 | hCoV-19/England/ALDP-B5BE2F/2020\|EPI_ISL_655710\|20 | ···················· | ···················· | ···················· | ···················· | ···················· | ···················· |
| S | 1148 | hCoV-19/England/QEUH-B6786E/2020\|EPI_ISL_655711\|20 | ···················· | ···················· | ···················· | ···················· | ···················· | ···················· |
| S | 1149 | hCoV-19/England/ALDP-B5C4A4/2020\|EPI_ISL_655712\|20 | ···················· | ···················· | ···················· | ···················· | ···················· | ···················· |
| S | 1150 | hCoV-19/England/ALDP-B5C2F5/2020\|EPI_ISL_655713\|20 | ···················· | ···················· | ···················· | ···················· | ···················· | ···················· |
| S | 1151 | hCoV-19/England/ALDP-B5C76F/2020\|EPI_ISL_655716\|20 | ···················· | ···················· | ···················· | ···················· | ···················· | ···················· |
| S | 1152 | hCoV-19/England/ALDP-B5BEC5/2020\|EPI_ISL_655718\|20 | ···················· | ···················· | ···················· | ···················· | ·····T·············· | ···················· |
| S | 1153 | hCoV-19/England/ALDP-B5C064/2020\|EPI_ISL_655720\|20 | ···················· | ·A·················· | ···················· | ···················· | ···················· | ··A················· |
| S | 1154 | hCoV-19/England/ALDP-B5BF3B/2020\|EPI_ISL_655721\|20 | ···················· | ···················· | ···················· | ···················· | ···················· | ···················· |
| S | 1155 | hCoV-19/England/ALDP-B5CEA6/2020\|EPI_ISL_655722\|20 | ···················· | ·A·················· | ···················· | ···················· | ···················· | ··A················· |
| S | 1156 | hCoV-19/England/MILK-B56B5A/2020\|EPI_ISL_656980\|20 | ···················· | ···················· | ···················· | ···················· | ···················· | ···················· |
| S | 1157 | hCoV-19/England/CAMC-B537FF/2020\|EPI_ISL_656981\|20 | ···················· | ···················· | ···················· | ···················· | ···················· | ···················· |
| S | 1158 | hCoV-19/England/MILK-B56817/2020\|EPI_ISL_656982\|20 | ···················· | ···················· | ···················· | ···················· | ·····T·············· | ···················· |
| S | 1159 | hCoV-19/England/CAMC-B54293/2020\|EPI_ISL_656983\|20 | ···················· | ···················· | ···················· | ···················· | ···················· | ···················· |
| S | 1160 | hCoV-19/England/CAMC-B522FF/2020\|EPI_ISL_656986\|20 | ···················· | ···················· | ···················· | ···················· | ···················· | ···················· |
| S | 1161 | hCoV-19/England/MILK-B56AE4/2020\|EPI_ISL_656987\|20 | ···················· | ···················· | ···················· | ···················· | ···················· | ···················· |
| S | 1162 | hCoV-19/England/CAMC-B54451/2020\|EPI_ISL_656999\|20 | ···················· | ···················· | ···················· | ···················· | ···················· | ···················· |
| S | 1163 | hCoV-19/England/CAMC-B33DF3/2020\|EPI_ISL_659200\|20 | ···················· | ···················· | ···················· | ···················· | ···················· | ···················· |
| S | 1164 | hCoV-19/England/CAMC-B32389/2020\|EPI_ISL_659201\|20 | ···················· | ···················· | ···················· | ···················· | ·····T·············· | ···················· |
| S | 1165 | hCoV-19/England/CAMC-B325DE/2020\|EPI_ISL_659203\|20 | ···················· | ···················· | ···················· | ···················· | ···················· | ···················· |
| S | 1166 | hCoV-19/England/CAMC-B339E0/2020\|EPI_ISL_659204\|20 | ···················· | ···················· | ···················· | ···················· | ···················· | ···················· |
| S | 1167 | hCoV-19/England/CAMC-B3244A/2020\|EPI_ISL_659205\|20 | ···················· | ·R·················· | ···················· | ···················· | ···················· | ···················· |
| S | 1168 | hCoV-19/England/CAMC-B33DE4/2020\|EPI_ISL_659206\|20 | ···················· | ···················· | ···················· | ···················· | ···················· | ···················· |
| S | 1169 | hCoV-19/England/CAMC-B33212/2020\|EPI_ISL_659208\|20 | ···················· | ···················· | ···················· | ···················· | ···················· | ···················· |
| S | 1170 | hCoV-19/England/CAMC-B321BC/2020\|EPI_ISL_659209\|20 | ···················· | ·A·················· | ···················· | ···················· | ···················· | ···················· |

| | | | | | | | |
|---|---|---|---|---|---|---|---|
| S | 1171 hCoV-19/England/CAMC-B33F75/2020\|EPI_ISL_659226\|20 | ···················· | ···················· | ····················· | ···················· | ···················· | ···················· |
| S | 1172 hCoV-19/England/CAMC-B32F2B/2020\|EPI_ISL_659253\|20 | ···················· | ···················· | ····················· | ···················· | ···················· | ···················· |
| S | 1173 hCoV-19/England/CAMC-B33F2A/2020\|EPI_ISL_659254\|20 | ···················· | ···················· | ····················· | ···················· | ···················· | ···················· |
| S | 1174 hCoV-19/England/CAMC-B333A6/2020\|EPI_ISL_659255\|20 | ···················· | ···················· | ····················· | ···················· | ·····T·············· | ···················· |
| S | 1175 hCoV-19/England/CAMC-B33E96/2020\|EPI_ISL_659257\|20 | ···················· | ···················· | ····················· | ···················· | ···················· | ···················· |
| S | 1176 hCoV-19/England/CAMC-B337C8/2020\|EPI_ISL_659260\|20 | ···················· | ···················· | ····················· | ···················· | ·····T·············· | ···················· |
| S | 1177 hCoV-19/England/CAMC-B325CF/2020\|EPI_ISL_659261\|20 | ···················· | ···················· | ····················· | ···················· | ···················· | ···················· |
| S | 1178 hCoV-19/England/CAMC-B33A56/2020\|EPI_ISL_659262\|20 | ···················· | ···················· | ····················· | ···················· | ···················· | ···················· |
| S | 1179 hCoV-19/England/CAMC-B33A83/2020\|EPI_ISL_659263\|20 | ···················· | ···················· | ····················· | ···················· | ·····T·············· | ···················· |
| S | 1180 hCoV-19/England/CAMC-B33DB7/2020\|EPI_ISL_659264\|20 | ···················· | ···················· | ····················· | ···················· | ···················· | ···················· |
| S | 1181 hCoV-19/England/CAMC-B33B71/2020\|EPI_ISL_659265\|20 | ···················· | ···················· | ····················· | ···················· | ···················· | ···················· |
| S | 1182 hCoV-19/England/CAMC-B3243B/2020\|EPI_ISL_659266\|20 | ···················· | ···················· | ····················· | ···················· | ···················· | ···················· |
| S | 1183 hCoV-19/England/CAMC-B338F2/2020\|EPI_ISL_659267\|20 | ···················· | ···················· | ····················· | ···················· | ···················· | ···················· |
| S | 1184 hCoV-19/England/CAMC-B33E2D/2020\|EPI_ISL_659268\|20 | ···················· | ···················· | ····················· | ···················· | ···················· | ···················· |
| S | 1185 hCoV-19/England/CAMC-B33D7B/2020\|EPI_ISL_659269\|20 | ···················· | ···················· | ····················· | ···················· | ···················· | ···················· |
| S | 1186 hCoV-19/England/CAMC-B3394A/2020\|EPI_ISL_659271\|20 | ···················· | ···················· | ····················· | ···················· | ···················· | ···················· |
| S | 1187 hCoV-19/England/CAMC-B32FD0/2020\|EPI_ISL_659272\|20 | ···················· | ···················· | ····················· | ···················· | ···················· | ···················· |
| S | 1188 hCoV-19/England/CAMC-B33151/2020\|EPI_ISL_659274\|20 | ···················· | ···················· | ····················· | ···················· | ···················· | ···················· |
| S | 1189 hCoV-19/England/CAMC-B33133/2020\|EPI_ISL_659275\|20 | ···················· | ···················· | ····················· | ···················· | ···················· | ···················· |
| S | 1190 hCoV-19/England/CAMC-B33027/2020\|EPI_ISL_659276\|20 | ···················· | ···················· | ····················· | ···················· | ·····T·············· | ···················· |
| S | 1191 hCoV-19/England/CAMC-B3329A/2020\|EPI_ISL_659277\|20 | ···················· | ···················· | ····················· | ···················· | ···················· | ···················· |
| S | 1192 hCoV-19/England/CAMC-B33BCC/2020\|EPI_ISL_659278\|20 | ···················· | ···················· | ····················· | ···················· | ···················· | ···················· |
| S | 1193 hCoV-19/England/CAMC-B33D6C/2020\|EPI_ISL_659279\|20 | ···················· | ···················· | ····················· | ···················· | ···················· | ···················· |
| S | 1194 hCoV-19/England/CAMC-B3240E/2020\|EPI_ISL_659280\|20 | ···················· | ···················· | ····················· | ···················· | ···················· | ···················· |
| S | 1195 hCoV-19/England/CAMC-B330CD/2020\|EPI_ISL_659281\|20 | ···················· | ···················· | ····················· | ···················· | ···················· | ···················· |
| S | 1196 hCoV-19/England/CAMC-B33397/2020\|EPI_ISL_659284\|20 | ···················· | ···················· | ····················· | ···················· | ···················· | ···················· |
| S | 1197 hCoV-19/England/CAMC-B32F1C/2020\|EPI_ISL_659285\|20 | ···················· | ···················· | ····················· | ···················· | ·····T·············· | ···················· |
| S | 1198 hCoV-19/England/CAMC-B32592/2020\|EPI_ISL_659286\|20 | ···················· | ···················· | ····················· | ···················· | ···················· | ···················· |
| S | 1199 hCoV-19/England/CAMC-B33124/2020\|EPI_ISL_659287\|20 | ···················· | ···················· | ····················· | ···················· | ···················· | ···················· |
| S | 1200 hCoV-19/England/CAMC-B33AA1/2020\|EPI_ISL_659288\|20 | ···················· | ···················· | ····················· | ···················· | ···················· | ···················· |
| S | 1201 hCoV-19/England/CAMC-B33A74/2020\|EPI_ISL_659289\|20 | ···················· | ·A·················· | ····················· | ···················· | ···················· | ··A················· |
| S | 1202 hCoV-19/England/CAMC-B333F1/2020\|EPI_ISL_659290\|20 | ···················· | ···················· | ····················· | ···················· | ···················· | ···················· |
| S | 1203 hCoV-19/England/CAMC-B32310/2020\|EPI_ISL_659291\|20 | ···················· | ···················· | ····················· | ···················· | ···················· | ···················· |
| S | 1204 hCoV-19/England/CAMC-B32E2E/2020\|EPI_ISL_659296\|20 | ···················· | ···················· | ····················· | ···················· | ···················· | ···················· |
| S | 1205 hCoV-19/England/CAMC-B33670/2020\|EPI_ISL_659297\|20 | ···················· | ···················· | ····················· | ···················· | ···················· | ···················· |
| S | 1206 hCoV-19/England/CAMC-B3229B/2020\|EPI_ISL_659298\|20 | ···················· | ···················· | ····················· | ···················· | ···················· | ···················· |
| S | 1207 hCoV-19/South Africa/KRISP-K004324/2020\|EPI_ISL_66 | ···················· | ·A·················· | ····················· | ···················· | ···················· | ···················· |
| S | 1208 hCoV-19/South Africa/KRISP-K004325/2020\|EPI_ISL_66 | ···················· | ···················· | ····················· | ···················· | ···················· | ···················· |
| S | 1209 hCoV-19/South Africa/KRISP-K004326/2020\|EPI_ISL_66 | ···················· | ·A·················· | ····················· | ···················· | ···················· | ···················· |
| S | 1210 hCoV-19/South Africa/KRISP-K004327/2020\|EPI_ISL_66 | ···················· | ···················· | ····················· | ···················· | ···················· | ···················· |
| S | 1211 hCoV-19/South Africa/KRISP-K004328/2020\|EPI_ISL_66 | ···················· | ···················· | ····················· | ···················· | ···················· | ···················· |
| S | 1212 hCoV-19/South Africa/KRISP-K004329/2020\|EPI_ISL_66 | ···················· | ···················· | ····················· | ···················· | ···················· | ···················· |
| S | 1213 hCoV-19/South Africa/KRISP-K004330/2020\|EPI_ISL_66 | ···················· | ···················· | ····················· | ···················· | ···················· | ···················· |
| S | 1214 hCoV-19/South Africa/KRISP-K004331/2020\|EPI_ISL_66 | ···················· | ·A·················· | ····················· | ···················· | ···················· | ···················· |
| S | 1215 hCoV-19/South Africa/KRISP-K004332/2020\|EPI_ISL_66 | ···················· | ···················· | ····················· | ···················· | ···················· | ···················· |
| S | 1216 hCoV-19/South Africa/KRISP-K004333/2020\|EPI_ISL_66 | ···················· | ·A·················· | ····················· | ···················· | ···················· | ···················· |
| S | 1217 hCoV-19/South Africa/KRISP-K004535-CS1/2020\|EPI_IS | ···················· | ···················· | ····················· | ···················· | ···················· | ···················· |
| S | 1218 hCoV-19/South Africa/KRISP-K004536/2020\|EPI_ISL_66 | ···················· | ···················· | ····················· | ···················· | ···················· | ···················· |
| S | 1219 hCoV-19/South Africa/KRISP-K004537/2020\|EPI_ISL_66 | ···················· | ···················· | ····················· | ···················· | ···················· | ···················· |
| S | 1220 hCoV-19/South Africa/KRISP-K004538/2020\|EPI_ISL_66 | ···················· | ···················· | ····················· | ···················· | ···················· | ···················· |
| S | 1221 hCoV-19/South Africa/KRISP-K004539/2020\|EPI_ISL_66 | ···················· | ···················· | ····················· | ···················· | ···················· | ···················· |
| S | 1222 hCoV-19/South Africa/KRISP-K004540/2020\|EPI_ISL_66 | ···················· | ···················· | ····················· | ···················· | ···················· | ···················· |
| S | 1223 hCoV-19/South Africa/KRISP-K004542/2020\|EPI_ISL_66 | ···················· | ···················· | ····················· | ···················· | ···················· | ···················· |
| S | 1224 hCoV-19/South Africa/KRISP-K004543/2020\|EPI_ISL_66 | ···················· | ···················· | ····················· | ···················· | ···················· | ···················· |
| S | 1225 hCoV-19/South Africa/KRISP-K004544/2020\|EPI_ISL_66 | ···················· | ···················· | ····················· | ···················· | ···················· | ···················· |
| S | 1226 hCoV-19/South Africa/KRISP-K004545/2020\|EPI_ISL_66 | ···················· | ···················· | ····················· | ···················· | ···················· | ···················· |
| S | 1227 hCoV-19/South Africa/KRISP-K004546/2020\|EPI_ISL_66 | ···················· | ···················· | ····················· | ···················· | ···················· | ···················· |
| S | 1228 hCoV-19/South Africa/KRISP-K004547/2020\|EPI_ISL_66 | ···················· | ···················· | ··T·················· | ···················· | ···················· | ···················· |
| S | 1229 hCoV-19/South Africa/KRISP-K004548/2020\|EPI_ISL_66 | ···················· | ···················· | ····················· | ···················· | ···················· | ···················· |
| S | 1230 hCoV-19/South Africa/KRISP-K004549/2020\|EPI_ISL_66 | ···················· | ···················· | ····················· | ···················· | ···················· | ···················· |
| S | 1231 hCoV-19/South Africa/KRISP-K004551/2020\|EPI_ISL_66 | ···················· | ···················· | ····················· | ···················· | ···················· | ···················· |
| S | 1232 hCoV-19/South Africa/KRISP-K004552/2020\|EPI_ISL_66 | ···················· | ···················· | ····················· | ···················· | ···················· | ···················· |
| S | 1233 hCoV-19/South Africa/KRISP-K004553/2020\|EPI_ISL_66 | ···················· | ···················· | ····················· | ···················· | ···················· | ···················· |
| S | 1234 hCoV-19/South Africa/KRISP-K004554/2020\|EPI_ISL_66 | ···················· | ···················· | ····················· | ···················· | ···················· | ···················· |
| S | 1235 hCoV-19/South Africa/KRISP-K004555/2020\|EPI_ISL_66 | ···················· | ···················· | ····················· | ···················· | ···················· | ···················· |
| S | 1236 hCoV-19/South Africa/KRISP-K004556/2020\|EPI_ISL_66 | ···················· | ···················· | ····················· | ···················· | ···················· | ···················· |
| S | 1237 hCoV-19/South Africa/KRISP-K004557/2020\|EPI_ISL_66 | ···················· | ···················· | ····················· | ···················· | ···················· | ···················· |
| S | 1238 hCoV-19/South Africa/KRISP-K004559/2020\|EPI_ISL_66 | ···················· | ···················· | ····················· | ···················· | ···················· | ···················· |
| S | 1239 hCoV-19/South Africa/KRISP-K004723-CS1/2020\|EPI_IS | ···················· | ···················· | ····················· | ···················· | ···················· | ···················· |
| S | 1240 hCoV-19/South Africa/KRISP-K004724-CS1/2020\|EPI_IS | ···················· | ···················· | ····················· | ···················· | ···················· | ···················· |
| S | 1241 hCoV-19/South Africa/KRISP-K004725-CS1/2020\|EPI_IS | ···················· | ···················· | ····················· | ···················· | ···················· | ···················· |
| S | 1242 hCoV-19/South Africa/KRISP-K004731/2020\|EPI_ISL_66 | ···················· | ···················· | ····················· | ···················· | ···················· | ···················· |
| S | 1243 hCoV-19/South Africa/KRISP-K004733/2020\|EPI_ISL_66 | ···················· | ·A·················· | ····················· | ···················· | ···················· | ···················· |
| S | 1244 hCoV-19/South Africa/KRISP-K004734/2020\|EPI_ISL_66 | ···················· | ···················· | ····················· | ···················· | ···················· | ···················· |
| S | 1245 hCoV-19/South Africa/KRISP-K004735/2020\|EPI_ISL_66 | ···················· | ···················· | ····················· | ···················· | ···················· | ···················· |
| S | 1246 hCoV-19/Spain/CT-IBV-99009868/2020\|EPI_ISL_660265\| | ···················· | ···················· | ····················· | ···················· | ···················· | ···················· |
| S | 1248 hCoV-19/Spain/CT-IBV-99009870/2020\|EPI_ISL_660267\| | ···················· | ···················· | ····················· | ···················· | ···················· | ···················· |
| S | 1249 hCoV-19/Spain/CT-IBV-99009871/2020\|EPI_ISL_660268\| | ···················· | ···················· | ····················· | ···················· | ···················· | ···················· |
| S | 1250 hCoV-19/Spain/CT-IBV-99009872/2020\|EPI_ISL_660269\| | ···················· | ···················· | ····················· | ···················· | ···················· | ···················· |
| S | 1251 hCoV-19/Spain/CT-IBV-99009873/2020\|EPI_ISL_660270\| | ···················· | ···················· | ····················· | ···················· | ·NN················· | ···················· |

```
S    1252 hCoV-19/Spain/CT-IBV-99009874/2020|EPI_ISL_660271|    ····················  ····················  ·····················  ····················  ····················  ····················
S    1253 hCoV-19/Spain/CT-IBV-99009875/2020|EPI_ISL_660272|    ····················  ····················  ·····················  ····················  ····················  ····················
S    1254 hCoV-19/Spain/CT-IBV-99009876/2020|EPI_ISL_660273|    ····················  ····················  ·····················  ····················  ····················  ····················
S    1255 hCoV-19/Spain/CT-IBV-99009877/2020|EPI_ISL_660274|    ····················  ····················  ·····················  ····················  ····················  ····················
S    1258 hCoV-19/Spain/CT-IBV-99009880/2020|EPI_ISL_660277|    ····················  ····················  ·····················  ····················  ····················  ····················
S    1259 hCoV-19/Spain/CT-IBV-99009881/2020|EPI_ISL_660278|    ····················  ····················  ·····················  ····················  ····················  ····················
S    1260 hCoV-19/Spain/CT-IBV-99009882/2020|EPI_ISL_660279|    ····················  ····················  ·····················  ····················  ····················  ····················
S    1261 hCoV-19/Spain/CT-IBV-99009883/2020|EPI_ISL_660280|    ····················  ····················  ·····················  ····················  ····················  ····················
S    1262 hCoV-19/Spain/CT-IBV-99009884/2020|EPI_ISL_660281|    ····················  ····················  ·····················  ····················  ····················  ····················
S    1263 hCoV-19/Spain/CT-IBV-99009885/2020|EPI_ISL_660282|    ····················  ····················  ·····················  ····················  ····················  ····················
S    1265 hCoV-19/Spain/CT-IBV-99009887/2020|EPI_ISL_660284|    ····················  ····················  ·····················  ····················  ····················  ····················
S    1266 hCoV-19/Spain/CT-IBV-99009888/2020|EPI_ISL_660285|    ····················  ····················  ·····················  ····················  ····················  ····················
S    1267 hCoV-19/Spain/CT-IBV-99009889/2020|EPI_ISL_660286|    ····················  ····················  ·····················  ····················  ····················  ····················
S    1268 hCoV-19/Spain/CT-IBV-99009890/2020|EPI_ISL_660287|    ····················  ····················  ·····················  ····················  ····················  ····················
S    1270 hCoV-19/Spain/CT-IBV-99009892/2020|EPI_ISL_660289|    ····················  ····················  ·····················  ····················  ····················  ····················
S    1272 hCoV-19/Spain/CT-IBV-99009894/2020|EPI_ISL_660291|    ····················  ····················  ·····················  ····················  ····················  ····················
S    1273 hCoV-19/Spain/CT-IBV-99009895/2020|EPI_ISL_660292|    ····················  ····················  ·····················  ····················  ····················  ····················
S    1274 hCoV-19/Spain/CT-IBV-99009896/2020|EPI_ISL_660293|    ····················  ····················  ·····················  ····················  ····················  ····················
S    1275 hCoV-19/Spain/CT-IBV-99009897/2020|EPI_ISL_660294|    ····················  ····················  ·····················  ····················  ····················  ····················
S    1276 hCoV-19/Spain/CT-IBV-99009898/2020|EPI_ISL_660295|    ····················  ····················  ·····················  ····················  ····················  ····················
S    1281 hCoV-19/England/C200042039/2020|EPI_ISL_649800|202    ····················  ····················  ·····················  ····················  ····················  ····················
S    1282 hCoV-19/England/C200042043/2020|EPI_ISL_649801|202    ····················  ····················  ·····················  ····················  ····················  ····················
S    1283 hCoV-19/England/C200042062/2020|EPI_ISL_649802|202    ····················  ····················  ·····················  ····················  ····················  ····················
S    1284 hCoV-19/England/C200042066/2020|EPI_ISL_649804|202    ····················  ····················  ·····················  ····················  ····················  ····················
S    1285 hCoV-19/England/C200042075/2020|EPI_ISL_649805|202    ····················  ····················  ·····················  ····················  ····················  ····················
S    1286 hCoV-19/England/C200042081/2020|EPI_ISL_649806|202    ····················  ····················  ·····················  ····················  ····················  ····················
S    1287 hCoV-19/England/C200042086/2020|EPI_ISL_649807|202    ····················  ····················  ·····················  ····················  ····················  ····················
S    1288 hCoV-19/England/204291476/2020|EPI_ISL_649810|2020    ····················  ····················  ·····················  ····················  ····················  ····················
S    1289 hCoV-19/England/204300490/2020|EPI_ISL_649812|2020    ····················  ·A··················  ·····················  ····················  ····················  ···A················
S    1290 hCoV-19/England/204300660/2020|EPI_ISL_649814|2020    ····················  ····················  ·····················  ····················  ····················  ····················
S    1291 hCoV-19/England/204300697/2020|EPI_ISL_649815|2020    ····················  ····················  ·····················  ····················  ····················  ····················
S    1292 hCoV-19/England/204300795/2020|EPI_ISL_649816|2020    ····················  ····················  ·····················  ····················  ····················  ····················
S    1293 hCoV-19/England/204300798/2020|EPI_ISL_649817|2020    ····················  ····················  ·····················  ····················  ············K·······  ····················
S    1294 hCoV-19/England/204320758/2020|EPI_ISL_649818|2020    ····················  ····················  ·····················  ····················  ····················  ····················
S    1295 hCoV-19/England/204320877/2020|EPI_ISL_649819|2020    ····················  ····················  ·····················  ····················  ····················  ····················
S    1296 hCoV-19/England/204320878/2020|EPI_ISL_649820|2020    ····················  ····················  ·····················  ····················  ····················  ····················
S    1297 hCoV-19/England/204321106/2020|EPI_ISL_649821|2020    ····················  ·A··················  ·····················  ····················  ····················  ···A················
S    1298 hCoV-19/England/204321118/2020|EPI_ISL_649822|2020    ····················  ····················  ·····················  ····················  ····················  ····················
S    1299 hCoV-19/England/204321284/2020|EPI_ISL_649823|2020    ····················  ····················  ·····················  ····················  ····················  ····················
S    1300 hCoV-19/England/204321411/2020|EPI_ISL_649824|2020    ····················  ····················  ·····················  ····················  ····················  ····················
S    1301 hCoV-19/England/204321417/2020|EPI_ISL_649825|2020    ····················  ·A··················  ·····················  ····················  ····················  ···A················
S    1302 hCoV-19/England/204321420/2020|EPI_ISL_649826|2020    ····················  ····················  ·····················  ····················  ····················  ····················
S    1303 hCoV-19/England/204321425/2020|EPI_ISL_649827|2020    ····················  ····················  ·····················  ····················  ····················  ····················
S    1304 hCoV-19/England/204321426/2020|EPI_ISL_649828|2020    ····················  ·A··················  ·····················  ····················  ····················  ···A················
S    1305 hCoV-19/England/204321427/2020|EPI_ISL_649829|2020    ····················  ·A··················  ·····················  ····················  ····················  ···A················
S    1306 hCoV-19/England/204341426/2020|EPI_ISL_649830|2020    ····················  ····················  ·····················  ····················  ····················  ····················
S    1307 hCoV-19/England/204341433/2020|EPI_ISL_649831|2020    ····················  ····················  ·····················  ····················  ····················  ····················
S    1308 hCoV-19/England/204341509/2020|EPI_ISL_649832|2020    ····················  ·A··················  ·····················  ····················  ····················  ···A················
S    1309 hCoV-19/England/204341522/2020|EPI_ISL_649833|2020    ····················  ·A··················  ·····················  ····················  ····················  ···A················
S    1310 hCoV-19/England/204341599/2020|EPI_ISL_649834|2020    ····················  ·A··················  ·····················  ····················  ····················  ···A················
S    1311 hCoV-19/England/204341622/2020|EPI_ISL_649838|2020    ····················  ····················  ·····················  ····················  ····················  ····················
S    1312 hCoV-19/England/204341624/2020|EPI_ISL_649839|2020    ····················  ····················  ·····················  ····················  ····················  ····················
S    1313 hCoV-19/England/204341629/2020|EPI_ISL_649840|2020    ····················  ····················  ·····················  ····················  ····················  ····················
S    1314 hCoV-19/England/204341652/2020|EPI_ISL_649841|2020    ····················  ····················  ·····················  ·············NN·····  ····················  ····················
S    1315 hCoV-19/England/204361024/2020|EPI_ISL_649842|2020    ····················  ····················  ·····················  ····················  ····················  ····················
S    1316 hCoV-19/England/204361148/2020|EPI_ISL_649843|2020    ····················  ····················  ·····················  ····················  ······T·············  ····················
S    1317 hCoV-19/England/204380383/2020|EPI_ISL_649844|2020    ····················  ····················  ·····················  ····················  ····················  ····················
S    1318 hCoV-19/England/204380844/2020|EPI_ISL_649846|2020    ····················  ·A··················  ·····················  ····················  ····················  ···A················
S    1319 hCoV-19/England/204380853/2020|EPI_ISL_649847|2020    ····················  ····················  ·····················  ····················  ····················  ····················
S    1320 hCoV-19/England/204380856/2020|EPI_ISL_649848|2020    ····················  ····················  ·····················  ····················  ····················  ····················
S    1321 hCoV-19/England/QEUH-AAED03/2020|EPI_ISL_653000|20    ····················  ····················  ·····················  ····················  ····················  ····················
S    1322 hCoV-19/England/QEUH-AAED21/2020|EPI_ISL_653001|20    ····················  ····················  ·····················  ····················  ····················  ····················
S    1323 hCoV-19/England/QEUH-AAED5E/2020|EPI_ISL_653002|20    ····················  ····················  ·····················  ····················  ····················  ····················
S    1324 hCoV-19/England/QEUH-AAED7C/2020|EPI_ISL_653003|20    ····················  ····················  ·····················  ····················  ····················  ····················
S    1325 hCoV-19/England/QEUH-AAED8B/2020|EPI_ISL_653004|20    ····················  ····················  ·····················  ····················  ····················  ····················
S    1326 hCoV-19/England/QEUH-AAED9A/2020|EPI_ISL_653005|20    ····················  ····················  ·····················  ····················  ····················  ····················
S    1327 hCoV-19/England/QEUH-AF5F07/2020|EPI_ISL_653030|20    ····················  ····················  ·····················  ····················  ····················  ····················
S    1328 hCoV-19/England/QEUH-AF5F16/2020|EPI_ISL_653031|20    ····················  ····················  ·····················  ····················  ····················  ····················
S    1329 hCoV-19/England/QEUH-AF5F25/2020|EPI_ISL_653032|20    ····················  ····················  ·····················  ····················  ····················  ····················
S    1330 hCoV-19/England/QEUH-AF5F34/2020|EPI_ISL_653033|20    ····················  ····················  ·····················  ····················  ····················  ····················
S    1331 hCoV-19/England/QEUH-AF5F43/2020|EPI_ISL_653034|20    ····················  ····················  ·····················  ····················  ····················  ····················
S    1332 hCoV-19/England/QEUH-AF5F8F/2020|EPI_ISL_653037|20    ····················  ····················  ·····················  ····················  ····················  ····················
S    1333 hCoV-19/England/QEUH-AF62D0/2020|EPI_ISL_653050|20    ····················  ····················  ·····················  ············A·······  ····················  ····················
S    1334 hCoV-19/England/QEUH-AF62EF/2020|EPI_ISL_653051|20    ····················  ····················  ·····················  ····················  ····················  ····················
S    1335 hCoV-19/England/QEUH-AF62FE/2020|EPI_ISL_653052|20    ····················  ····················  ·····················  ····················  ····················  ····················
S    1336 hCoV-19/England/QEUH-AF630A/2020|EPI_ISL_653053|20    ····················  ····················  ·····················  ····················  ····················  ····················
S    1337 hCoV-19/Scotland/QEUH-AF6346/2020|EPI_ISL_653054|2    ····················  ····················  ·····················  ····················  ····················  ····················
S    1338 hCoV-19/England/QEUH-AF63A0/2020|EPI_ISL_653055|20    ····················  ····················  ·····················  ····················  ····················  ····················
S    1339 hCoV-19/England/QEUH-AF63BF/2020|EPI_ISL_653056|20    ····················  ····················  ·····················  ····················  ····················  ····················
S    1340 hCoV-19/England/QEUH-AF63CE/2020|EPI_ISL_653057|20    ····················  ····················  ·····················  ····················  ····················  ····················
```

| | | | | | | | |
|---|---|---|---|---|---|---|---|
| S | 1341 hCoV-19/Wales/QEUH-AF63DD/2020\|EPI_ISL_653058\|2020 | ···················· | ···················· | ····················· | ···················· | ···················· | ···················· |
| S | 1342 hCoV-19/England/SHEF-C0B38/2020\|EPI_ISL_653059\|202 | ···················· | ···················· | ····················· | ···················· | ···················· | ···················· |
| S | 1343 hCoV-19/England/SHEF-C0E4E/2020\|EPI_ISL_653060\|202 | ···················· | ···················· | ····················· | ···················· | ···················· | ···················· |
| S | 1344 hCoV-19/England/SHEF-C2538/2020\|EPI_ISL_653061\|202 | ···················· | ···················· | ····················· | ···················· | ···················· | ···················· |
| S | 1345 hCoV-19/England/SHEF-C334C/2020\|EPI_ISL_653062\|202 | ···················· | ···················· | ····················· | ···················· | ···················· | ···················· |
| S | 1346 hCoV-19/England/SHEF-C3591/2020\|EPI_ISL_653063\|202 | ···················· | ·A·················· | ····················· | ···················· | ···················· | ···················· |
| S | 1347 hCoV-19/England/SHEF-C3704/2020\|EPI_ISL_653064\|202 | ···················· | ···················· | ····················· | ···················· | ···················· | ···················· |
| S | 1348 hCoV-19/England/SHEF-C3BBD/2020\|EPI_ISL_653065\|202 | ···················· | ···················· | ····················· | ···················· | ···················· | ···················· |
| S | 1349 hCoV-19/England/SHEF-C3BEA/2020\|EPI_ISL_653066\|202 | ···················· | ···················· | ····················· | ···················· | ···················· | ···················· |
| S | 1350 hCoV-19/England/SHEF-C4F38/2020\|EPI_ISL_653067\|202 | ···················· | ···················· | ····················· | ···················· | ···················· | ···················· |
| S | 1351 hCoV-19/England/SHEF-C53D1/2020\|EPI_ISL_653068\|202 | ···················· | ···················· | ····················· | ···················· | ···················· | ···················· |
| S | 1352 hCoV-19/England/SHEF-C56D8/2020\|EPI_ISL_653069\|202 | ···················· | ···················· | ····················· | ···················· | ···················· | ···················· |
| S | 1353 hCoV-19/England/SHEF-C675C/2020\|EPI_ISL_653070\|202 | ···················· | ···················· | ····················· | ···················· | ···················· | ···················· |
| S | 1354 hCoV-19/England/SHEF-C6F81/2020\|EPI_ISL_653071\|202 | ···················· | ···················· | ····················· | ···················· | ···················· | ···················· |
| S | 1355 hCoV-19/England/SHEF-C7BE6/2020\|EPI_ISL_653072\|202 | ···················· | ···················· | ····················· | ···················· | ···················· | ···················· |
| S | 1356 hCoV-19/England/SHEF-C8356/2020\|EPI_ISL_653073\|202 | ···················· | ···················· | ····················· | ···················· | ···················· | ···················· |
| S | 1357 hCoV-19/England/SHEF-C9337/2020\|EPI_ISL_653074\|202 | ···················· | ···················· | ····················· | ···················· | ···················· | ···················· |
| S | 1358 hCoV-19/England/SHEF-C964D/2020\|EPI_ISL_653075\|202 | ···················· | ·A·················· | ····················· | ···················· | ···················· | ···················· |
| S | 1359 hCoV-19/England/SHEF-CB818/2020\|EPI_ISL_653076\|202 | ···················· | ···················· | ····················· | ···················· | ···················· | ···················· |
| S | 1360 hCoV-19/England/SHEF-CB942/2020\|EPI_ISL_653077\|202 | ···················· | ···················· | ····················· | ···················· | ···················· | ···················· |
| S | 1361 hCoV-19/England/SHEF-CBCB2/2020\|EPI_ISL_653078\|202 | ···················· | ···················· | ····················· | ···················· | ···················· | ···················· |
| S | 1362 hCoV-19/England/SHEF-CBD28/2020\|EPI_ISL_653079\|202 | ···················· | ···················· | ····················· | ···················· | ···················· | ···················· |
| S | 1363 hCoV-19/England/SHEF-CBD91/2020\|EPI_ISL_653080\|202 | ···················· | ···················· | ····················· | ···················· | ···················· | ···················· |
| S | 1364 hCoV-19/England/SHEF-CC352/2020\|EPI_ISL_653081\|202 | ···················· | ···················· | ····················· | ···················· | ···················· | ···················· |
| S | 1365 hCoV-19/England/SHEF-CD764/2020\|EPI_ISL_653082\|202 | ···················· | ···················· | ····················· | ···················· | ···················· | ···················· |
| S | 1366 hCoV-19/England/SHEF-CDA01/2020\|EPI_ISL_653083\|202 | ···················· | ···················· | ····················· | ···················· | ···················· | ···················· |
| S | 1367 hCoV-19/England/SHEF-CDBD1/2020\|EPI_ISL_653084\|202 | ···················· | ·A·················· | ····················· | ···················· | ···················· | ···················· |
| S | 1368 hCoV-19/England/SHEF-CE3AB/2020\|EPI_ISL_653085\|202 | ···················· | ···················· | ····················· | ···················· | ···················· | ···················· |
| S | 1369 hCoV-19/England/SHEF-CE4A8/2020\|EPI_ISL_653086\|202 | ···················· | ···················· | ····················· | ···················· | ···················· | ···················· |
| S | 1370 hCoV-19/England/SHEF-CE53C/2020\|EPI_ISL_653087\|202 | ···················· | ···················· | ····················· | ···················· | ···················· | ···················· |
| S | 1371 hCoV-19/England/SHEF-CE824/2020\|EPI_ISL_653088\|202 | ···················· | ···················· | ····················· | ···················· | ···················· | ···················· |
| S | 1372 hCoV-19/England/SHEF-CEEB9/2020\|EPI_ISL_653089\|202 | ···················· | ···················· | ····················· | ···················· | ···················· | ···················· |
| S | 1373 hCoV-19/England/SHEF-CEEC8/2020\|EPI_ISL_653090\|202 | ···················· | ···················· | ····················· | ···················· | ···················· | ···················· |
| S | 1374 hCoV-19/England/SHEF-CF1BF/2020\|EPI_ISL_653091\|202 | ···················· | ···················· | ····················· | ···················· | ···················· | ···················· |
| S | 1375 hCoV-19/England/SHEF-CF2F8/2020\|EPI_ISL_653092\|202 | ···················· | ···················· | ····················· | ···················· | ···················· | ···················· |
| S | 1376 hCoV-19/England/SHEF-D13E1/2020\|EPI_ISL_653093\|202 | ···················· | ···················· | ····················· | ···················· | ···················· | ···················· |
| S | 1377 hCoV-19/England/SHEF-D1C5F/2020\|EPI_ISL_653094\|202 | ···················· | ···················· | ····················· | ···················· | ···················· | ···················· |
| S | 1378 hCoV-19/England/SHEF-D2D3D/2020\|EPI_ISL_653095\|202 | ···················· | ···················· | ····················· | ···················· | ···················· | ···················· |
| S | 1379 hCoV-19/England/SHEF-D31F4/2020\|EPI_ISL_653096\|202 | ···················· | ···················· | ····················· | ···················· | ···················· | ···················· |
| S | 1380 hCoV-19/England/SHEF-D3200/2020\|EPI_ISL_653097\|202 | ···················· | ···················· | ····················· | ···················· | ···················· | ···················· |
| S | 1381 hCoV-19/USA/FL-BPHL-1945/2020\|EPI_ISL_653098\|2020- | ···················· | ···················· | ····················· | ···················· | ···················· | ···················· |
| S | 1383 hCoV-19/Spain/MD-IBV-99008728/2020\|EPI_ISL_654301\| | ···················· | ···················· | ····················· | ···················· | ···················· | ···················· |
| S | 1393 hCoV-19/Spain/MD-IBV-99008749/2020\|EPI_ISL_654311\| | ···················· | ···················· | ····················· | ···················· | ···················· | ···················· |
| S | 1401 hCoV-19/Spain/MD-IBV-99008776/2020\|EPI_ISL_654319\| | ···················· | ···················· | ····················· | ···················· | ···················· | ···················· |
| S | 1405 hCoV-19/Spain/MD-IBV-99009619/2020\|EPI_ISL_654336\| | ···················· | ···················· | ····················· | ···················· | ···················· | ···················· |
| S | 1408 hCoV-19/Spain/MD-IBV-99009620/2020\|EPI_ISL_654362\| | ···················· | ···················· | ····················· | ···················· | ···················· | ···················· |
| S | 1412 hCoV-19/Spain/MD-IBV-99009628/2020\|EPI_ISL_654368\| | ···················· | ···················· | ····················· | ···················· | ···················· | ···················· |
| S | 1416 hCoV-19/Spain/MD-IBV-99009648/2020\|EPI_ISL_654382\| | ···················· | ···················· | ····················· | ···················· | ···················· | ···················· |
| S | 1432 hCoV-19/Spain/MD-IBV-99009673/2020\|EPI_ISL_654398\| | ···················· | ···················· | ····················· | ···················· | ···················· | ···················· |
| S | 1433 hCoV-19/Spain/MD-IBV-99009674/2020\|EPI_ISL_654399\| | ···················· | ···················· | ····················· | ···················· | ···················· | ···················· |
| S | 1434 hCoV-19/England/ALDP-B62D73/2020\|EPI_ISL_655602\|20 | ···················· | ···················· | ····················· | ···················· | ···················· | ···················· |
| S | 1435 hCoV-19/England/ALDP-B5BB55/2020\|EPI_ISL_655603\|20 | ···················· | ···················· | ····················· | ···················· | ···················· | ···················· |
| S | 1436 hCoV-19/England/ALDP-B5E69D/2020\|EPI_ISL_655605\|20 | ···················· | ···················· | ····················· | ···················· | ···················· | ···················· |
| S | 1437 hCoV-19/England/ALDP-B5AF69/2020\|EPI_ISL_655607\|20 | ···················· | ···················· | ····················· | ···················· | ···················· | ···················· |
| S | 1438 hCoV-19/England/ALDP-B5BAD0/2020\|EPI_ISL_655608\|20 | ···················· | ···················· | ····················· | ···················· | ···················· | ···················· |
| S | 1439 hCoV-19/England/ALDP-B62EDA/2020\|EPI_ISL_655609\|20 | ···················· | ···················· | ····················· | ···················· | ···················· | ···················· |
| S | 1440 hCoV-19/England/ALDP-B5BC52/2020\|EPI_ISL_655612\|20 | ···················· | ···················· | ····················· | ···················· | ···················· | ···················· |
| S | 1441 hCoV-19/England/ALDP-B5B487/2020\|EPI_ISL_655613\|20 | ···················· | ···················· | ····················· | ···················· | ···················· | ···················· |
| S | 1442 hCoV-19/England/ALDP-B5E6F7/2020\|EPI_ISL_655614\|20 | ···················· | ···················· | ····················· | ···················· | ···················· | ···················· |
| S | 1443 hCoV-19/England/ALDP-B62C94/2020\|EPI_ISL_655615\|20 | ···················· | ···················· | ····················· | ···················· | ···················· | ···················· |
| S | 1444 hCoV-19/England/ALDP-B5E6BB/2020\|EPI_ISL_655616\|20 | ···················· | ···················· | ····················· | ···················· | ···················· | ···················· |
| S | 1445 hCoV-19/England/ALDP-B63C48/2020\|EPI_ISL_655617\|20 | ···················· | ···················· | ····················· | ···················· | ···················· | ···················· |
| S | 1446 hCoV-19/England/ALDP-B5BD31/2020\|EPI_ISL_655619\|20 | ···················· | ···················· | ····················· | ···················· | ···················· | ···················· |
| S | 1447 hCoV-19/England/ALDP-B5E730/2020\|EPI_ISL_655620\|20 | ···················· | ···················· | ····················· | ···················· | ···················· | ···················· |
| S | 1448 hCoV-19/England/ALDP-B5E651/2020\|EPI_ISL_655622\|20 | ···················· | ···················· | ····················· | ···················· | ···················· | ···················· |
| S | 1449 hCoV-19/England/ALDP-B5BD6E/2020\|EPI_ISL_655623\|20 | ···················· | ···················· | ····················· | ···················· | ···················· | ···················· |
| S | 1450 hCoV-19/England/ALDP-B5B85E/2020\|EPI_ISL_655624\|20 | ···················· | ···················· | ····················· | ···················· | ···················· | ···················· |
| S | 1451 hCoV-19/England/ALDP-B5B52A/2020\|EPI_ISL_655625\|20 | ···················· | ···················· | ····················· | ···················· | ···················· | ···················· |
| S | 1452 hCoV-19/England/ALDP-B62C76/2020\|EPI_ISL_655626\|20 | ···················· | ···················· | ····················· | ···················· | ···················· | ···················· |
| S | 1453 hCoV-19/England/ALDP-B62E9E/2020\|EPI_ISL_655628\|20 | ···················· | ···················· | ····················· | ···················· | ···················· | ···················· |
| S | 1454 hCoV-19/England/ALDP-B5B5EE/2020\|EPI_ISL_655630\|20 | ···················· | ···················· | ····················· | ···················· | ···················· | ···················· |
| S | 1455 hCoV-19/England/ALDP-B5B478/2020\|EPI_ISL_655631\|20 | ···················· | ···················· | ····················· | ···················· | ···················· | ···················· |
| S | 1456 hCoV-19/England/ALDP-B5AFD2/2020\|EPI_ISL_655632\|20 | ···················· | ···················· | ····················· | ···················· | ···················· | ···················· |
| S | 1457 hCoV-19/England/ALDP-B5B997/2020\|EPI_ISL_655633\|20 | ···················· | ···················· | ····················· | ···················· | ···················· | ···················· |
| S | 1458 hCoV-19/England/ALDP-B62D19/2020\|EPI_ISL_655634\|20 | ···················· | ···················· | ····················· | ···················· | ···················· | ···················· |
| S | 1459 hCoV-19/England/ALDP-B5B2E7/2020\|EPI_ISL_655635\|20 | ···················· | ···················· | ····················· | ···················· | ···················· | ···················· |
| S | 1460 hCoV-19/England/ALDP-B5B1DB/2020\|EPI_ISL_655636\|20 | ···················· | ···················· | ····················· | ···················· | ···················· | ···················· |
| S | 1461 hCoV-19/England/ALDP-B62D82/2020\|EPI_ISL_655637\|20 | ···················· | ···················· | ····················· | ···················· | ···················· | ···················· |
| S | 1462 hCoV-19/England/ALDP-B5CC8E/2020\|EPI_ISL_655638\|20 | ···················· | ···················· | ····················· | ···················· | ···················· | ···················· |
| S | 1463 hCoV-19/England/ALDP-B5C705/2020\|EPI_ISL_655640\|20 | ···················· | ···················· | ····················· | ···················· | ···················· | ···················· |

S 1464 hCoV-19/England/ALDP-B5C44A/2020|EPI_ISL_655641|20 ···················· ···················· ····················· ···················· ···················· ····················
S 1465 hCoV-19/England/ALDP-B5BF77/2020|EPI_ISL_655642|20 ···················· ···················· ····················· ···················· ······T············· ····················
S 1466 hCoV-19/England/ALDP-B5C529/2020|EPI_ISL_655644|20 ···················· ···················· ····················· ···················· ···················· ····················
S 1467 hCoV-19/England/ALDP-B5C134/2020|EPI_ISL_655645|20 ···················· ·A·················· ····················· ···················· ···················· ···A················
S 1468 hCoV-19/England/ALDP-B5C26E/2020|EPI_ISL_655646|20 ···················· ···················· ····················· ···················· ···················· ····················
S 1469 hCoV-19/England/ALDP-B5BDD7/2020|EPI_ISL_655649|20 ···················· ···················· ····················· ···················· ···················· ····················
S 1470 hCoV-19/England/ALDP-B5C41D/2020|EPI_ISL_655658|20 ···················· ···················· ····················· ···················· ···················· ····················
S 1471 hCoV-19/England/ALDP-B5C802/2020|EPI_ISL_655659|20 ···················· ·········T·········· ····················· ···················· ···················· ····················
S 1472 hCoV-19/England/QEUH-B677F8/2020|EPI_ISL_655660|20 ···················· ···················· ····················· ···················· ···················· ····················
S 1473 hCoV-19/England/ALDP-B5C899/2020|EPI_ISL_655661|20 ···················· ···················· ····················· ···················· ···················· ····················
S 1474 hCoV-19/England/QEUH-B65F2F/2020|EPI_ISL_655662|20 ···················· ···················· ····················· ···················· ···················· ····················
S 1475 hCoV-19/England/ALDP-B5CAFD/2020|EPI_ISL_655663|20 ···················· ···················· ····················· ···················· ···················· ····················
S 1476 hCoV-19/England/ALDP-B5C6DB/2020|EPI_ISL_655664|20 ···················· ···················· ····················· ···················· ···················· ····················
S 1477 hCoV-19/England/ALDP-B5CC9D/2020|EPI_ISL_655665|20 ···················· ···················· ····················· ···················· ···················· ····················
S 1478 hCoV-19/England/ALDP-B5BE10/2020|EPI_ISL_655666|20 ···················· ···················· ····················· ···················· ···················· ····················
S 1479 hCoV-19/England/ALDP-B5CE97/2020|EPI_ISL_655667|20 ···················· ···················· ····················· ···················· ···················· ····················
S 1480 hCoV-19/England/ALDP-B5C4C2/2020|EPI_ISL_655690|20 ···················· ···················· ····················· ···················· ···················· ····················
S 1481 hCoV-19/England/ALDP-B5CBFA/2020|EPI_ISL_655691|20 ···················· ···················· ····················· ···················· ······T············· ····················
S 1482 hCoV-19/England/ALDP-B5BE01/2020|EPI_ISL_655692|20 ···················· ···················· ····················· ···················· ···················· ····················
S 1483 hCoV-19/England/ALDP-B5C3E3/2020|EPI_ISL_655693|20 ···················· ···················· ····················· ···················· ···················· ····················
S 1484 hCoV-19/England/QEUH-B6788C/2020|EPI_ISL_655694|20 ···················· ···················· ····················· ···················· ···················· ····················
S 1485 hCoV-19/England/QEUH-B65E6E/2020|EPI_ISL_655695|20 ···················· ···················· ····················· ···················· ···················· ····················
S 1486 hCoV-19/England/QEUH-B65F6B/2020|EPI_ISL_655699|20 ···················· ···················· ····················· ···················· ···················· ····················
S 1487 hCoV-19/England/CAMC-B54381/2020|EPI_ISL_656933|20 ···················· ···················· ····················· ···················· ···················· ····················
S 1488 hCoV-19/England/CAMC-B53391/2020|EPI_ISL_656934|20 ···················· ···················· ····················· ···················· ···················· ····················
S 1489 hCoV-19/England/MILK-B569BA/2020|EPI_ISL_656935|20 ···················· ···················· ····················· ···················· ···················· ····················
S 1490 hCoV-19/England/MILK-B56CFD/2020|EPI_ISL_656936|20 ···················· ···················· ····················· ···················· ···················· ····················
S 1491 hCoV-19/England/CAMC-B53FAC/2020|EPI_ISL_656937|20 ···················· ····A··············· ····················· ···················· ···················· ····················
S 1492 hCoV-19/England/CAMC-B54390/2020|EPI_ISL_656940|20 ···················· ···················· ····················· ···················· ······T············· ····················
S 1493 hCoV-19/England/CAMC-B5220E/2020|EPI_ISL_656941|20 ···················· ···················· ····················· ···················· ···················· ····················
S 1494 hCoV-19/England/MILK-B56871/2020|EPI_ISL_656942|20 ···················· ···················· ····················· ···················· ···················· ····················
S 1495 hCoV-19/England/CAMC-B5410F/2020|EPI_ISL_656943|20 ···················· ···················· ····················· ···················· ···················· ····················
S 1496 hCoV-19/England/CAMC-B54011/2020|EPI_ISL_656944|20 ···················· ···················· ····················· ···················· ···················· ····················
S 1497 hCoV-19/England/MILK-B56844/2020|EPI_ISL_656945|20 ···················· ···················· ····················· ···················· ···················· ····················
S 1498 hCoV-19/England/MILK-B56941/2020|EPI_ISL_656947|20 ···················· ···················· ····················· ···················· ···················· ····················
S 1499 hCoV-19/England/CAMC-B54372/2020|EPI_ISL_656949|20 ···················· ···················· ····················· ···················· ······T············· ····················
S 1500 hCoV-19/England/MILK-B56C48/2020|EPI_ISL_656950|20 ···················· ·A·················· ····················· ···················· ···················· ···A················
S 1501 hCoV-19/England/CAMC-B52541/2020|EPI_ISL_656952|20 ···················· ···················· ····················· ···················· ···················· ····················
S 1502 hCoV-19/England/CAMC-B52602/2020|EPI_ISL_656953|20 ···················· ···················· ····················· ···················· ···················· ····················
S 1503 hCoV-19/England/CAMC-B533DD/2020|EPI_ISL_656954|20 ···················· ···················· ····················· ···················· ···················· ····················
S 1504 hCoV-19/England/MILK-B56AF3/2020|EPI_ISL_656955|20 ···················· ···················· ····················· ···················· ···················· ····················
S 1505 hCoV-19/England/CAMC-B52365/2020|EPI_ISL_656956|20 ···················· ···················· ····················· ···················· ···················· ····················
S 1506 hCoV-19/England/MILK-B568DB/2020|EPI_ISL_656957|20 ···················· ···················· ····················· ···················· ···················· ····················
S 1507 hCoV-19/England/MILK-B56A8A/2020|EPI_ISL_656958|20 ···················· ···················· ····················· ···················· ···················· ····················
S 1508 hCoV-19/England/CAMC-B53777/2020|EPI_ISL_656959|20 ···················· ···················· ····················· ···················· ···················· ····················
S 1509 hCoV-19/England/CAMC-B54178/2020|EPI_ISL_656960|20 ···················· ···················· ····················· ···················· ···················· ····················
S 1510 hCoV-19/England/MILK-B568EA/2020|EPI_ISL_656961|20 ···················· ···················· ····················· ···················· ···················· ····················
S 1511 hCoV-19/England/MILK-B56923/2020|EPI_ISL_656962|20 ···················· ·A·················· ····················· ···················· ···················· ···A················
S 1512 hCoV-19/England/CAMC-B53935/2020|EPI_ISL_656963|20 ···················· ···················· ····················· ···················· ······T············· ····················
S 1513 hCoV-19/England/CAMC-B54363/2020|EPI_ISL_656964|20 ···················· ···················· ····················· ···················· ···················· ····················
S 1514 hCoV-19/England/MILK-B56774/2020|EPI_ISL_656965|20 ···················· ···················· ····················· ···················· ···················· ····················
S 1515 hCoV-19/England/MILK-B56A6C/2020|EPI_ISL_656966|20 ···················· ···················· ····················· ···················· ···················· ····················
S 1516 hCoV-19/England/CAMC-B53856/2020|EPI_ISL_656967|20 ···················· ···················· ····················· ···················· ···················· ····················
S 1517 hCoV-19/England/CAMC-B523A1/2020|EPI_ISL_656968|20 ···················· ·A·················· ····················· ···················· ···················· ···A················
S 1518 hCoV-19/England/MILK-B56C0C/2020|EPI_ISL_656969|20 ···················· ···················· ····················· ···················· ···················· ····················
S 1519 hCoV-19/England/CAMC-B52329/2020|EPI_ISL_656970|20 ···················· ···················· ····················· ···················· ···················· ····················
S 1520 hCoV-19/England/CAMC-B53892/2020|EPI_ISL_656971|20 ···················· ·A·················· ····················· ···················· ···················· ···A················
S 1521 hCoV-19/England/CAMC-B53944/2020|EPI_ISL_656972|20 ···················· ···················· ····················· ···················· ···················· ····················
S 1522 hCoV-19/England/MILK-B56932/2020|EPI_ISL_656973|20 ···················· ···················· ····················· ···················· ······T············· ····················
S 1523 hCoV-19/England/MILK-B5699C/2020|EPI_ISL_656974|20 ···················· ·A·················· ····················· ···················· ···················· ···A················
S 1524 hCoV-19/England/CAMC-B53847/2020|EPI_ISL_656975|20 ···················· ···················· ····················· ···················· ···················· ····················
S 1525 hCoV-19/England/MILK-B56862/2020|EPI_ISL_656976|20 ···················· ···················· ····················· ···················· ······T············· ····················
S 1526 hCoV-19/England/CAMC-B53452/2020|EPI_ISL_656977|20 ···················· ···················· ····················· ···················· ···················· ····················
S 1527 hCoV-19/England/CAMC-B5271E/2020|EPI_ISL_656978|20 ···················· ···················· ····················· ···················· ···················· ····················
S 1528 hCoV-19/England/MILK-B568F9/2020|EPI_ISL_656979|20 ···················· ···················· ····················· ···················· ···················· ····················
S 1529 hCoV-19/England/CAMC-B53FD9/2020|EPI_ISL_656989|20 ···················· ···················· ····················· ···················· ···················· ····················
S 1530 hCoV-19/England/CAMC-B54275/2020|EPI_ISL_656990|20 ···················· ···················· ····················· ···················· ···················· ····················
S 1531 hCoV-19/England/MILK-B56A3F/2020|EPI_ISL_656991|20 ···················· ···················· ····················· ···················· ···················· ····················
S 1532 hCoV-19/England/CAMC-B541E1/2020|EPI_ISL_656992|20 ···················· ·A·················· ····················· ···················· ···················· ····················
S 1533 hCoV-19/England/CAMC-B5449D/2020|EPI_ISL_656993|20 ···················· ···················· ····················· ···················· ···················· ····················
S 1534 hCoV-19/England/CAMC-B523FC/2020|EPI_ISL_656995|20 ···················· ···················· ····················· ···················· ···················· ····················
S 1535 hCoV-19/England/MILK-B5697E/2020|EPI_ISL_656997|20 ···················· ···················· ····················· ···················· ···················· ····················
S 1536 hCoV-19/England/CAMC-B53FBB/2020|EPI_ISL_656998|20 ···················· ····A··············· ····················· ···················· ···················· ····················
S 1537 hCoV-19/Scotland/QEUH-AD0500/2020|EPI_ISL_649728|2 ···················· ···················· ····················· ···················· ···················· ····················
S 1538 hCoV-19/Scotland/QEUH-AD668F/2020|EPI_ISL_649729|2 ···················· ···················· ····················· ···················· ···················· ····················
S 1539 hCoV-19/Scotland/QEUH-AD4144/2020|EPI_ISL_649730|2 ···················· ···················· ····················· ···················· ···················· ····················
S 1540 hCoV-19/Scotland/QEUH-AD0209/2020|EPI_ISL_649731|2 ···················· ···················· ····················· ···················· ···················· ····················
S 1541 hCoV-19/Scotland/QEUH-AD0175/2020|EPI_ISL_649733|2 ···················· ···················· ····················· ···················· ···················· ····················
S 1542 hCoV-19/Scotland/QEUH-AD051F/2020|EPI_ISL_649734|2 ···················· ···················· ····················· ···················· ···················· ····················
S 1543 hCoV-19/Scotland/QEUH-AD2182/2020|EPI_ISL_649735|2 ···················· ···················· ····················· ···················· ···················· ····················

| | | | | | | | | |
|---|---|---|---|---|---|---|---|---|
| S | 1544 hCoV-19/Scotland/QEUH-AD2119/2020\|EPI_ISL_649736\|2 | ···················· | ···················· | ···················· | ···················· | ···················· | ···················· | |
| S | 1545 hCoV-19/Scotland/QEUH-AD01DF/2020\|EPI_ISL_649737\|2 | ···················· | ···················· | ···················· | ···················· | ···················· | ···················· | |
| S | 1546 hCoV-19/Scotland/QEUH-AD2173/2020\|EPI_ISL_649738\|2 | ···················· | ···················· | ···················· | ···················· | ···················· | ···················· | |
| S | 1547 hCoV-19/Scotland/QEUH-AD6670/2020\|EPI_ISL_649739\|2 | ···················· | ···················· | ···················· | ···················· | ···················· | ···················· | |
| S | 1548 hCoV-19/Scotland/QEUH-AD21DD/2020\|EPI_ISL_649741\|2 | ···················· | ···················· | ···················· | ···················· | ···················· | ···················· | |
| S | 1549 hCoV-19/Scotland/QEUH-AD66E9/2020\|EPI_ISL_649742\|2 | ···················· | ···················· | ···················· | ···················· | ···················· | ···················· | |
| S | 1550 hCoV-19/Scotland/QEUH-AD6722/2020\|EPI_ISL_649743\|2 | ···················· | ···················· | ···················· | ···················· | ···················· | ···················· | |
| S | 1551 hCoV-19/Scotland/QEUH-AD2128/2020\|EPI_ISL_649745\|2 | ···················· | ···················· | ···················· | ···················· | ···················· | ···················· | |
| S | 1552 hCoV-19/Scotland/QEUH-AD0C29/2020\|EPI_ISL_649747\|2 | ···················· | ···················· | ···················· | ···················· | ···················· | ···················· | |
| S | 1553 hCoV-19/Scotland/QEUH-AD20FE/2020\|EPI_ISL_649748\|2 | ···················· | ···················· | ···················· | ···················· | ···················· | ···················· | |
| S | 1554 hCoV-19/Scotland/QEUH-AD0597/2020\|EPI_ISL_649749\|2 | ···················· | ···················· | ···················· | ···················· | ···················· | ···················· | |
| S | 1555 hCoV-19/Scotland/QEUH-AD2137/2020\|EPI_ISL_649752\|2 | ···················· | ···················· | ···················· | ···················· | ···················· | ···················· | |
| S | 1556 hCoV-19/Scotland/QEUH-AD492E/2020\|EPI_ISL_649753\|2 | ···················· | ···················· | ···················· | ···················· | ···················· | ···················· | |
| S | 1557 hCoV-19/Scotland/QEUH-AD66DA/2020\|EPI_ISL_649754\|2 | ···················· | ···················· | ···················· | ···················· | ···················· | ···················· | |
| S | 1558 hCoV-19/Scotland/QEUH-AD0227/2020\|EPI_ISL_649755\|2 | ···················· | ···················· | ···················· | ···················· | ···················· | ···················· | |
| S | 1559 hCoV-19/Scotland/QEUH-AD66CB/2020\|EPI_ISL_649757\|2 | ···················· | ···················· | ···················· | ···················· | ···················· | ···················· | |
| S | 1560 hCoV-19/Scotland/QEUH-AD6661/2020\|EPI_ISL_649758\|2 | ···················· | ···················· | ···················· | ···················· | ···················· | ···················· | |
| S | 1561 hCoV-19/Scotland/QEUH-AD0184/2020\|EPI_ISL_649759\|2 | ···················· | ···················· | ···················· | ···················· | ···················· | ···················· | |
| S | 1562 hCoV-19/Scotland/QEUH-AD2191/2020\|EPI_ISL_649760\|2 | ···················· | ···················· | ···················· | ···················· | ···················· | ···················· | |
| S | 1563 hCoV-19/Scotland/QEUH-AD01FD/2020\|EPI_ISL_649761\|2 | ···················· | ···················· | ···················· | ···················· | ···················· | ···················· | |
| S | 1564 hCoV-19/Scotland/QEUH-AD052E/2020\|EPI_ISL_649762\|2 | ···················· | ···················· | ···················· | ···················· | ···················· | ···················· | |
| S | 1565 hCoV-19/Scotland/QEUH-AD0218/2020\|EPI_ISL_649763\|2 | ···················· | ···················· | ···················· | ···················· | ···················· | ···················· | |
| S | 1566 hCoV-19/Scotland/QEUH-AD01C0/2020\|EPI_ISL_649764\|2 | ···················· | ···················· | ···················· | ···················· | ···················· | ···················· | |
| S | 1567 hCoV-19/Scotland/QEUH-AD6731/2020\|EPI_ISL_649765\|2 | ···················· | ···················· | ···················· | ···················· | ···················· | ···················· | |
| S | 1568 hCoV-19/Scotland/QEUH-AD21BF/2020\|EPI_ISL_649767\|2 | ···················· | ···················· | ···················· | ···················· | ···················· | ···················· | |
| S | 1569 hCoV-19/Scotland/QEUH-AD0254/2020\|EPI_ISL_649768\|2 | ···················· | ···················· | ···················· | ···················· | ···················· | ···················· | |
| S | 1570 hCoV-19/Scotland/QEUH-AD05B5/2020\|EPI_ISL_649769\|2 | ···················· | ···················· | ···················· | ···················· | ···················· | ···················· | |
| S | 1571 hCoV-19/Scotland/QEUH-AD0245/2020\|EPI_ISL_649770\|2 | ···················· | ···················· | ···················· | ···················· | ···················· | ···················· | |
| S | 1572 hCoV-19/Scotland/QEUH-AD6740/2020\|EPI_ISL_649771\|2 | ···················· | ···················· | ···················· | ···················· | ···················· | ···················· | |
| S | 1573 hCoV-19/Scotland/QEUH-AD2155/2020\|EPI_ISL_649772\|2 | ···················· | ···················· | ···················· | ···················· | ···················· | ···················· | |
| S | 1574 hCoV-19/Scotland/QEUH-AD2094/2020\|EPI_ISL_649773\|2 | ···················· | ···················· | ···················· | ···················· | ···················· | ···················· | |
| S | 1575 hCoV-19/Scotland/QEUH-AD04F4/2020\|EPI_ISL_649774\|2 | ···················· | ···················· | ···················· | ···················· | ···················· | ···················· | |
| S | 1576 hCoV-19/Scotland/QEUH-AD05F1/2020\|EPI_ISL_649775\|2 | ···················· | ···················· | ···················· | ···················· | ···················· | ···················· | |
| S | 1577 hCoV-19/Scotland/QEUH-AD0579/2020\|EPI_ISL_649776\|2 | ···················· | ···················· | ···················· | ···················· | ···················· | ···················· | |
| S | 1578 hCoV-19/Scotland/QEUH-AD0773/2020\|EPI_ISL_649777\|2 | ···················· | ···················· | ···················· | ···················· | ···················· | ···················· | |
| S | 1579 hCoV-19/Scotland/QEUH-AD2049/2020\|EPI_ISL_649778\|2 | ···················· | ···················· | ···················· | ···················· | ···················· | ···················· | |
| S | 1580 hCoV-19/Scotland/QEUH-AD07FB/2020\|EPI_ISL_649779\|2 | ···················· | ···················· | ···················· | ···················· | ···················· | ···················· | |
| S | 1581 hCoV-19/Scotland/QEUH-AD6704/2020\|EPI_ISL_649780\|2 | ···················· | ···················· | ···················· | ···················· | ···················· | ···················· | |
| S | 1582 hCoV-19/Scotland/QEUH-AD21A0/2020\|EPI_ISL_649781\|2 | ···················· | ···················· | ···················· | ···················· | ···················· | ···················· | |
| S | 1583 hCoV-19/Scotland/QEUH-AD66AD/2020\|EPI_ISL_649782\|2 | ···················· | ···················· | ···················· | ···················· | ···················· | ···················· | |
| S | 1584 hCoV-19/Scotland/QEUH-AD055B/2020\|EPI_ISL_649783\|2 | ···················· | ···················· | ···················· | ···················· | ···················· | ···················· | |
| S | 1585 hCoV-19/Italy/BAS-IZSPB-173APT/2020\|EPI_ISL_649784 | ···················· | ···················· | ···················· | ···················· | ···················· | ···················· | |
| S | 1586 hCoV-19/Italy/APU-IZSPB-182PT/2020\|EPI_ISL_649785\| | ···················· | ···················· | ···················· | ···················· | ···················· | ···················· | |
| S | 1587 hCoV-19/England/C200041963/2020\|EPI_ISL_649786\|202 | ···················· | ···················· | ···················· | ···················· | ···················· | ···················· | |
| S | 1588 hCoV-19/England/C200041964/2020\|EPI_ISL_649787\|202 | ···················· | ···················· | ···················· | ···················· | ···················· | ···················· | |
| S | 1589 hCoV-19/England/C200041972/2020\|EPI_ISL_649788\|202 | ···················· | ···················· | ···················· | ···················· | ···················· | ···················· | |
| S | 1590 hCoV-19/England/C200041976/2020\|EPI_ISL_649789\|202 | ···················· | ···················· | ···················· | ···················· | ···················· | ···················· | |
| S | 1591 hCoV-19/England/C200041985/2020\|EPI_ISL_649791\|202 | ···················· | ···················· | ···················· | ···················· | ···················· | ···················· | |
| S | 1592 hCoV-19/England/C200042006/2020\|EPI_ISL_649793\|202 | ···················· | ···················· | ···················· | ···················· | ···················· | ···················· | |
| S | 1593 hCoV-19/England/C200042014/2020\|EPI_ISL_649794\|202 | ···················· | ···················· | ···················· | ···················· | ···················· | ···················· | |
| S | 1594 hCoV-19/England/C200042020/2020\|EPI_ISL_649795\|202 | ···················· | ···················· | ···················· | ···················· | ···················· | ···················· | |
| S | 1595 hCoV-19/England/C200042027/2020\|EPI_ISL_649796\|202 | ···················· | ···················· | ···················· | ···················· | ···················· | ···················· | |
| S | 1596 hCoV-19/England/C200042029/2020\|EPI_ISL_649797\|202 | ···················· | ···················· | ···················· | ···················· | ···················· | ···················· | |
| S | 1597 hCoV-19/England/C200042030/2020\|EPI_ISL_649798\|202 | ···················· | ···················· | ···················· | ···················· | ···················· | ···················· | |
| S | 1598 hCoV-19/England/C200042031/2020\|EPI_ISL_649799\|202 | ···················· | ···················· | ···················· | ···················· | ···················· | ···················· | |
| S | 1599 hCoV-19/England/QEUH-AF556F/2020\|EPI_ISL_653028\|20 | ···················· | ·A·················· | ···················· | ···················· | ···················· | ···················· | |
| S | 1600 hCoV-19/England/QEUH-AF55F6/2020\|EPI_ISL_653029\|20 | ···················· | ···················· | ···················· | ···················· | ···················· | ···················· | |
| S | 1601 hCoV-19/England/QEUH-AF5F52/2020\|EPI_ISL_653035\|20 | ···················· | ···················· | ···················· | ···················· | ···················· | ···················· | |
| S | 1602 hCoV-19/England/QEUH-AF5F61/2020\|EPI_ISL_653036\|20 | ···················· | ···················· | ···················· | ···················· | ···················· | ···················· | |
| S | 1603 hCoV-19/England/QEUH-AF5F9E/2020\|EPI_ISL_653038\|20 | ···················· | ···················· | ···················· | ···················· | ···················· | ···················· | |
| S | 1604 hCoV-19/England/QEUH-AF5FAD/2020\|EPI_ISL_653039\|20 | ···················· | ···················· | ···················· | ···················· | ···················· | ···················· | |
| S | 1605 hCoV-19/England/QEUH-AF5FBC/2020\|EPI_ISL_653040\|20 | ···················· | ···················· | ···················· | ···················· | ···················· | ···················· | |
| S | 1606 hCoV-19/England/QEUH-AF5FE9/2020\|EPI_ISL_653041\|20 | ···················· | ···················· | ···················· | ···················· | ···················· | ···················· | |
| S | 1607 hCoV-19/England/QEUH-AF6012/2020\|EPI_ISL_653042\|20 | ···················· | ···················· | ···················· | ···················· | ···················· | ···················· | |
| S | 1608 hCoV-19/England/QEUH-AF604F/2020\|EPI_ISL_653043\|20 | ··T················· | ···················· | ···················· | ···················· | ···················· | ···················· | |
| S | 1609 hCoV-19/England/QEUH-AF606D/2020\|EPI_ISL_653044\|20 | ···················· | ···················· | ···················· | ···················· | ···················· | ···················· | |
| S | 1610 hCoV-19/England/QEUH-AF60E5/2020\|EPI_ISL_653045\|20 | ···················· | ···················· | ···················· | ···················· | ···················· | ···················· | |
| S | 1611 hCoV-19/England/QEUH-AF6267/2020\|EPI_ISL_653046\|20 | ···················· | ···················· | ···················· | ···················· | ···················· | ···················· | |
| S | 1612 hCoV-19/England/QEUH-AF6285/2020\|EPI_ISL_653047\|20 | ···················· | ···················· | ···················· | ···················· | ···················· | ···················· | |
| S | 1613 hCoV-19/England/QEUH-AF62A3/2020\|EPI_ISL_653048\|20 | ···················· | ···················· | ···················· | ···················· | ···················· | ···················· | |
| S | 1614 hCoV-19/Wales/QEUH-AF62B2/2020\|EPI_ISL_653049\|2020 | ···················· | ···················· | ···················· | ···················· | ···················· | ···················· | |
| S | 1616 hCoV-19/Spain/MD-IBV-99008660/2020\|EPI_ISL_654221\| | ···················· | ···················· | ···················· | ···················· | ···················· | ···················· | |
| S | 1617 hCoV-19/Spain/MD-IBV-99008661/2020\|EPI_ISL_654222\| | ···················· | ···················· | ···················· | ···················· | ·NNN················ | ···················· | 1 |
| S | 1632 hCoV-19/Spain/MD-IBV-99009644/2020\|EPI_ISL_654379\| | ···················· | ···················· | ···················· | ···················· | ···················· | ···················· | |
| S | 1633 hCoV-19/England/ALDP-B5B36C/2020\|EPI_ISL_655551\|20 | ···················· | ···················· | ···················· | ···················· | ···················· | ···················· | |
| S | 1634 hCoV-19/England/ALDP-B5AFE1/2020\|EPI_ISL_655552\|20 | ···················· | ···················· | ···················· | ···················· | ···················· | ···················· | |
| S | 1635 hCoV-19/England/ALDP-B5B7D9/2020\|EPI_ISL_655553\|20 | ···················· | ···················· | ···················· | ···················· | ···················· | ···················· | |
| S | 1636 hCoV-19/England/ALDP-B5AEC6/2020\|EPI_ISL_655555\|20 | ···················· | ···················· | ···················· | ···················· | ···················· | ···················· | |
| S | 1637 hCoV-19/England/ALDP-B5AEA8/2020\|EPI_ISL_655556\|20 | ···················· | ···················· | ···················· | ···················· | ···················· | ···················· | |
| S | 1638 hCoV-19/England/ALDP-B5B6DC/2020\|EPI_ISL_655557\|20 | ···················· | ···················· | ···················· | ···················· | ···················· | ···················· | |

S 1639 hCoV-19/England/ALDP-B5B0DE/2020|EPI_ISL_655601|20 ···················· ···················· ···················· ···················· ···················· ····················
S 1640 hCoV-19/England/ALDP-B5C2AA/2020|EPI_ISL_655698|20 ···················· ···················· ···················· ···················· ···················· ····················
S 1641 hCoV-19/England/CAMC-B535F5/2020|EPI_ISL_656880|20 ···················· ···················· ···················· ···················· ···················· ····················
S 1642 hCoV-19/England/CAMC-B53829/2020|EPI_ISL_656882|20 ···················· ···················· ···················· ···················· ···················· ····················
S 1643 hCoV-19/England/CAMC-B526F3/2020|EPI_ISL_656883|20 ···················· ···················· ···················· ···················· ···················· ····················
S 1644 hCoV-19/England/CAMC-B525E7/2020|EPI_ISL_656885|20 ···················· ···················· ···················· ···················· ···················· ····················
S 1645 hCoV-19/England/CAMC-B52277/2020|EPI_ISL_656886|20 ···················· ···················· ···················· ···················· ···················· ····················
S 1646 hCoV-19/England/MILK-B56AB7/2020|EPI_ISL_656887|20 ···················· ···················· ···················· ···················· ···················· ····················
S 1647 hCoV-19/England/CAMC-B525AB/2020|EPI_ISL_656900|20 ···················· ···················· ···················· ···················· ···················· ····················
S 1648 hCoV-19/England/CAMC-B54406/2020|EPI_ISL_656902|20 ···················· ···················· ···················· ···················· ······T············· ····················
S 1649 hCoV-19/England/CAMC-B5447F/2020|EPI_ISL_656903|20 ···················· ···················· ···················· ···················· ···················· ····················
S 1650 hCoV-19/England/CAMC-B534AD/2020|EPI_ISL_656904|20 ···················· ···················· ···················· ···················· ···················· ····················
S 1651 hCoV-19/England/CAMC-B53434/2020|EPI_ISL_656905|20 ···················· ···················· ···················· ···················· ···················· ····················
S 1652 hCoV-19/England/MILK-B569C9/2020|EPI_ISL_656906|20 ···················· ···················· ···················· ···················· ···················· ····················
S 1653 hCoV-19/England/CAMC-B5407B/2020|EPI_ISL_656907|20 ···················· ···················· ···················· ···················· ···················· ····················
S 1654 hCoV-19/England/CAMC-B53953/2020|EPI_ISL_656909|20 ···················· ···················· ···················· ···················· ···················· ····················
S 1655 hCoV-19/England/MILK-B56B69/2020|EPI_ISL_656910|20 ···················· ···················· ···················· ···················· ···················· ····················
S 1656 hCoV-19/England/CAMC-B5256F/2020|EPI_ISL_656911|20 ···················· ···················· ···················· ···················· ···················· ····················
S 1657 hCoV-19/England/CAMC-B521C5/2020|EPI_ISL_656912|20 ···················· ···················· ···················· ···················· ···················· ····················
S 1658 hCoV-19/England/MILK-B568CC/2020|EPI_ISL_656913|20 ···················· ···················· ···················· ···················· ···················· ····················
S 1659 hCoV-19/England/CAMC-B54503/2020|EPI_ISL_656914|20 ···················· ···················· ···················· ···················· ···················· ····················
S 1660 hCoV-19/England/CAMC-B537C2/2020|EPI_ISL_656916|20 ···················· ···················· ···················· ···················· ···················· ····················
S 1661 hCoV-19/England/CAMC-B5270F/2020|EPI_ISL_656917|20 ···················· ·A·······A·········· ···················· ···················· ···················· ····················
S 1662 hCoV-19/England/CAMC-B535B9/2020|EPI_ISL_656918|20 ···················· ···················· ···················· ···················· ···················· ····················
S 1663 hCoV-19/England/CAMC-B5370E/2020|EPI_ISL_656919|20 ···················· ···················· ···················· ···················· ···················· ····················
S 1664 hCoV-19/England/CAMC-B52417/2020|EPI_ISL_656921|20 ···················· ···················· ···················· ···················· ···················· ····················
S 1665 hCoV-19/England/MILK-B568BD/2020|EPI_ISL_656922|20 ···················· ···················· ···················· ···················· ···················· ····················
S 1666 hCoV-19/England/CAMC-B53A23/2020|EPI_ISL_656923|20 ···················· ···················· ···················· ···················· ······T············· ····················
S 1667 hCoV-19/England/MILK-B5689F/2020|EPI_ISL_656924|20 ···················· ···················· ···················· ···················· ···················· ····················
S 1668 hCoV-19/England/CAMC-B53F60/2020|EPI_ISL_656925|20 ···················· ···················· ···················· ···················· ···················· ····················
S 1669 hCoV-19/England/CAMC-B5357D/2020|EPI_ISL_656926|20 ···················· ···················· ···················· ···················· ···················· ····················
S 1670 hCoV-19/England/CAMC-B54318/2020|EPI_ISL_656928|20 ···················· ···················· ···················· ···················· ···················· ····················
S 1671 hCoV-19/England/CAMC-B523CF/2020|EPI_ISL_656929|20 ···················· ···················· ···················· ···················· ···················· ····················
S 1672 hCoV-19/England/MILK-B56914/2020|EPI_ISL_656930|20 ···················· ···················· ···················· ···················· ···················· ····················
S 1673 hCoV-19/England/CAMC-B54257/2020|EPI_ISL_656931|20 ···················· ···················· ···················· ···················· ···················· ····················
S 1674 hCoV-19/England/MILK-B56B96/2020|EPI_ISL_656932|20 ···················· ···················· ···················· ···················· ······T············· ····················
S 1675 hCoV-19/England/CAMC-B32301/2020|EPI_ISL_659100|20 ···················· ·A·······A·········· ···················· ···················· ···················· ····················
S 1676 hCoV-19/England/CAMC-B33634/2020|EPI_ISL_659102|20 ···················· ···················· ···················· ···················· ···················· ····················
S 1677 hCoV-19/England/CAMC-B32BFA/2020|EPI_ISL_659105|20 ···················· ···················· ···················· ···················· ···················· ····················
S 1678 hCoV-19/England/CAMC-B33009/2020|EPI_ISL_659107|20 ···················· ···················· ···················· ···················· ···················· ····················
S 1679 hCoV-19/England/CAMC-B3375F/2020|EPI_ISL_659109|20 ···················· ···················· ···················· ···················· ···················· ····················
S 1680 hCoV-19/England/CAMC-B33740/2020|EPI_ISL_659110|20 ···················· ·········A·········· ···················· ···················· ······T············· ····················
S 1681 hCoV-19/England/CAMC-B33EE1/2020|EPI_ISL_659111|20 ···················· ···················· ···················· ···················· ···················· ····················
S 1682 hCoV-19/England/CAMC-B336AD/2020|EPI_ISL_659112|20 ···················· ···················· ···················· ···················· ···················· ····················
S 1683 hCoV-19/England/CAMC-B324A4/2020|EPI_ISL_659113|20 ···················· ···················· ···················· ···················· ···················· ····················
S 1684 hCoV-19/England/CAMC-B3393B/2020|EPI_ISL_659114|20 ···················· ···················· ···················· ···················· ···················· ····················
S 1685 hCoV-19/England/CAMC-B33476/2020|EPI_ISL_659115|20 ···················· ···················· ···················· ···················· ···················· ····················
S 1686 hCoV-19/England/CAMC-B33898/2020|EPI_ISL_659116|20 ···················· ···················· ···················· ···················· ···················· ····················
S 1687 hCoV-19/England/CAMC-B337E6/2020|EPI_ISL_659117|20 ···················· ···················· ···················· ···················· ···················· ····················
S 1688 hCoV-19/England/CAMC-B3227D/2020|EPI_ISL_659119|20 ···················· ···················· ···················· ···················· ···················· ····················
S 1689 hCoV-19/England/CAMC-B321CB/2020|EPI_ISL_659120|20 ···················· ···················· ···················· ···················· ···················· ····················
S 1690 hCoV-19/England/CAMC-B33995/2020|EPI_ISL_659121|20 ···················· ···················· ···················· ···················· ···················· ····················
S 1691 hCoV-19/England/CAMC-B323D4/2020|EPI_ISL_659122|20 ···················· ···················· ···················· ···················· ···················· ····················
S 1692 hCoV-19/England/CAMC-B325A1/2020|EPI_ISL_659125|20 ···················· ·A·················· ···················· ···················· ···················· ····················
S 1693 hCoV-19/England/CAMC-B337B9/2020|EPI_ISL_659126|20 ···················· ···················· ···················· ···················· ···················· ····················
S 1694 hCoV-19/England/CAMC-B337F5/2020|EPI_ISL_659130|20 ···················· ···················· ···················· ···················· ···················· ····················
S 1695 hCoV-19/England/CAMC-B334C1/2020|EPI_ISL_659131|20 ···················· ·A·················· ···················· ···················· ···················· ···A················
S 1696 hCoV-19/England/CAMC-B33BEA/2020|EPI_ISL_659133|20 ···················· ···················· ···················· ···················· ···················· ····················
S 1697 hCoV-19/England/CAMC-B321DA/2020|EPI_ISL_659134|20 ···················· ···················· ···················· ···················· ···················· ····················
S 1698 hCoV-19/England/CAMC-B334A3/2020|EPI_ISL_659135|20 ···················· ···················· ···················· ···················· ···················· ····················
S 1699 hCoV-19/England/CAMC-B33713/2020|EPI_ISL_659136|20 ···················· ·A·················· ···················· ···················· ···················· ···A················
S 1700 hCoV-19/England/CAMC-B33731/2020|EPI_ISL_659137|20 ···················· ···················· ···················· ···················· ···················· ····················
S 1701 hCoV-19/England/CAMC-B33582/2020|EPI_ISL_659138|20 ···················· ···················· ···················· ···················· ···················· ····················
S 1702 hCoV-19/England/CAMC-B332A9/2020|EPI_ISL_659141|20 ···················· ···················· ···················· ···················· ···················· ····················
S 1703 hCoV-19/England/CAMC-B332F4/2020|EPI_ISL_659142|20 ···················· ···················· ···················· ···················· ···················· ····················
S 1704 hCoV-19/England/CAMC-B336BC/2020|EPI_ISL_659144|20 ···················· ···················· ···················· ···················· ···················· ····················
S 1705 hCoV-19/England/CAMC-B338B6/2020|EPI_ISL_659145|20 ···················· ···················· ···················· ···················· ···················· ····················
S 1706 hCoV-19/England/CAMC-B33494/2020|EPI_ISL_659148|20 ···················· ···················· ···················· ···················· ···················· ····················
S 1707 hCoV-19/England/CAMC-B322F5/2020|EPI_ISL_659149|20 ···················· ·A·······A·········· ···················· ···················· ···················· ····················
S 1708 hCoV-19/England/CAMC-B33054/2020|EPI_ISL_659150|20 ···················· ···················· ···················· ···················· ···················· ····················
S 1709 hCoV-19/England/CAMC-B32231/2020|EPI_ISL_659151|20 ···················· ···················· ···················· ···················· ···················· ····················
S 1710 hCoV-19/England/CAMC-B33564/2020|EPI_ISL_659153|20 ···················· ···················· ···················· ···················· ···················· ····················
S 1711 hCoV-19/England/CAMC-B3378C/2020|EPI_ISL_659155|20 ···················· ···················· ···················· ···················· ···················· ····················
S 1712 hCoV-19/England/CAMC-B3383E/2020|EPI_ISL_659159|20 ···················· ···················· ···················· ···················· ···················· ····················
S 1713 hCoV-19/England/CAMC-B33968/2020|EPI_ISL_659160|20 ···················· ···················· ···················· ···················· ···················· ····················
S 1714 hCoV-19/England/CAMC-B33555/2020|EPI_ISL_659162|20 ···················· ···················· ···················· ···················· ···················· ····················
S 1715 hCoV-19/England/CAMC-B334D0/2020|EPI_ISL_659163|20 ···················· ···················· ···················· ···················· ···················· ····················
S 1716 hCoV-19/England/CAMC-B33FEE/2020|EPI_ISL_659164|20 ···················· ···················· ···················· ···················· ···················· ····················
S 1717 hCoV-19/England/CAMC-B33B17/2020|EPI_ISL_659165|20 ···················· ·A·················· ···················· ···················· ···················· ···A················
S 1718 hCoV-19/England/CAMC-B33D5D/2020|EPI_ISL_659166|20 ···················· ···················· ···················· ···················· ···················· ····················

| | | | | | | | |
|---|---|---|---|---|---|---|---|
| S | 1719 hCoV-19/England/CAMC-B32222/2020\|EPI_ISL_659168\|20 | ···················· | ···················· | ····················· | ···················· | ···················· | ···················· |
| S | 1720 hCoV-19/England/CAMC-B33537/2020\|EPI_ISL_659169\|20 | ···················· | ···················· | ····················· | ···················· | ···················· | ···················· |
| S | 1721 hCoV-19/England/CAMC-B33A0B/2020\|EPI_ISL_659171\|20 | ···················· | ···················· | ····················· | ···················· | ···················· | ···················· |
| S | 1722 hCoV-19/England/CAMC-B32459/2020\|EPI_ISL_659174\|20 | ···················· | ···················· | ····················· | ···················· | ······T············· | ···················· |
| S | 1723 hCoV-19/England/CAMC-B325FC/2020\|EPI_ISL_659175\|20 | ···················· | ···················· | ····················· | ···················· | ···················· | ···················· |
| S | 1724 hCoV-19/England/CAMC-B322B9/2020\|EPI_ISL_659177\|20 | ···················· | ···················· | ····················· | ···················· | ···················· | ···················· |
| S | 1725 hCoV-19/England/CAMC-B32C06/2020\|EPI_ISL_659179\|20 | ···················· | ···················· | ····················· | ···················· | ···················· | ···················· |
| S | 1726 hCoV-19/England/CAMC-B337AA/2020\|EPI_ISL_659180\|20 | ···················· | ···················· | ····················· | ···················· | ···················· | ······T············· |
| S | 1727 hCoV-19/England/CAMC-B324B3/2020\|EPI_ISL_659181\|20 | ···················· | ···················· | ····················· | ···················· | ···················· | ···················· |
| S | 1728 hCoV-19/England/CAMC-B32529/2020\|EPI_ISL_659182\|20 | ···················· | ···················· | ····················· | ···················· | ···················· | ···················· |
| S | 1729 hCoV-19/England/CAMC-B32662/2020\|EPI_ISL_659183\|20 | ···················· | ···················· | ····················· | ···················· | ···················· | ···················· |
| S | 1730 hCoV-19/England/CAMC-B3386B/2020\|EPI_ISL_659185\|20 | ···················· | ···················· | ····················· | ···················· | ···················· | ···················· |
| S | 1731 hCoV-19/England/CAMC-B3336A/2020\|EPI_ISL_659186\|20 | ···················· | ···················· | ····················· | ···················· | ···················· | ···················· |
| S | 1732 hCoV-19/England/CAMC-B333E2/2020\|EPI_ISL_659187\|20 | ···················· | ···················· | ····················· | ···················· | ···················· | ···················· |
| S | 1733 hCoV-19/England/CAMC-B33B44/2020\|EPI_ISL_659189\|20 | ···················· | ···················· | ····················· | ···················· | ···················· | ···················· |
| S | 1734 hCoV-19/England/CAMC-B32583/2020\|EPI_ISL_659190\|20 | ···················· | ·A·················· | ····················· | ···················· | ···················· | ···················· |
| S | 1735 hCoV-19/England/CAMC-B32E97/2020\|EPI_ISL_659192\|20 | ···················· | ···················· | ····················· | ···················· | ···················· | ···················· |
| S | 1736 hCoV-19/England/CAMC-B33A38/2020\|EPI_ISL_659193\|20 | ···················· | ···················· | ····················· | ···················· | ···················· | ···················· |
| S | 1737 hCoV-19/England/CAMC-B331AC/2020\|EPI_ISL_659194\|20 | ···················· | ·A·················· | ····················· | ···················· | ···················· | ···A················ |
| S | 1738 hCoV-19/England/CAMC-B33388/2020\|EPI_ISL_659195\|20 | ···················· | ···················· | ····················· | ···················· | ···················· | ···················· |
| S | 1739 hCoV-19/England/CAMC-B33E0F/2020\|EPI_ISL_659196\|20 | ···················· | ···················· | ····················· | ···················· | ···················· | ···················· |
| S | 1740 hCoV-19/England/CAMC-B33B53/2020\|EPI_ISL_659198\|20 | ···················· | ···················· | ····················· | ···················· | ···················· | ···················· |
| S | 1741 hCoV-19/USA/MD-MDH-0377/2020\|EPI_ISL_660100\|2020-1 | ···················· | ···················· | ····················· | ···················· | ···················· | ···················· |
| S | 1742 hCoV-19/USA/MD-MDH-0378/2020\|EPI_ISL_660101\|2020-1 | ···················· | ···················· | ····················· | ···················· | ···················· | ···················· |
| S | 1743 hCoV-19/USA/MD-MDH-0379/2020\|EPI_ISL_660102\|2020-1 | ···················· | ···················· | ····················· | ···················· | ···················· | ···················· |
| S | 1744 hCoV-19/USA/MD-MDH-0404/2020\|EPI_ISL_660103\|2020-1 | ···················· | ···················· | ····················· | ···················· | ···················· | ···················· |
| S | 1745 hCoV-19/USA/MD-MDH-0386/2020\|EPI_ISL_660104\|2020-1 | ···················· | ···················· | ····················· | ···················· | ···················· | ···················· |
| S | 1746 hCoV-19/USA/MD-MDH-0387/2020\|EPI_ISL_660105\|2020-1 | ···················· | ···················· | ····················· | ···················· | ···················· | ···················· |
| S | 1747 hCoV-19/South Korea/YS006/2020\|EPI_ISL_660109\|2020 | ···················· | ···················· | ··T·················· | ···················· | ···················· | ···················· |
| S | 1748 hCoV-19/South Korea/YS008/2020\|EPI_ISL_660110\|2020 | ···················· | ···················· | ····················· | ···················· | ···················· | ···················· |
| S | 1749 hCoV-19/South Africa/Tygerberg_312_CC2/2020\|EPI_IS | ···················· | ···················· | ····················· | ···················· | ···················· | ···················· |
| S | 1750 hCoV-19/South Africa/Tygerberg_313_CC2/2020\|EPI_IS | ···················· | ···················· | ····················· | ···················· | ···················· | ···················· |
| S | 1751 hCoV-19/South Africa/Tygerberg_315_CC2/2020\|EPI_IS | ···················· | ···················· | ····················· | ···················· | ···················· | ···················· |
| S | 1752 hCoV-19/South Africa/Tygerberg_318_CC2/2020\|EPI_IS | ···················· | ·A·················· | ····················· | ···················· | ···················· | ···················· |
| S | 1753 hCoV-19/South Africa/Tygerberg_331_CC2/2020\|EPI_IS | ···················· | ···················· | ····················· | ···················· | ···················· | ···················· |
| S | 1754 hCoV-19/South Africa/Tygerberg_334_CC2/2020\|EPI_IS | ···················· | ···················· | ····················· | ···················· | ···················· | ···················· |
| S | 1755 hCoV-19/South Africa/Tygerberg_336_CC2/2020\|EPI_IS | ···················· | ···················· | ····················· | ···················· | ···················· | ···················· |
| S | 1756 hCoV-19/South Africa/Tygerberg_337_CC2/2020\|EPI_IS | ···················· | ·A·················· | ····················· | ···················· | ···················· | ···················· |
| S | 1757 hCoV-19/South Africa/Tygerberg_338_CC2/2020\|EPI_IS | ···················· | ···················· | ····················· | ···················· | ···················· | ···················· |
| S | 1758 hCoV-19/South Africa/Tygerberg_339_CC2/2020\|EPI_IS | ···················· | ···················· | ····················· | ···················· | ···················· | ···················· |
| S | 1759 hCoV-19/South Africa/Tygerberg_344_CC2/2020\|EPI_IS | ···················· | ···················· | ····················· | ···················· | ···················· | ···················· |
| S | 1760 hCoV-19/South Africa/Tygerberg_346_CC2/2020\|EPI_IS | ···················· | ···················· | ····················· | ···················· | ···················· | ···················· |
| S | 1761 hCoV-19/South Africa/Tygerberg_349_CC2/2020\|EPI_IS | ···················· | ···················· | ····················· | ···················· | ···················· | ···················· |
| S | 1762 hCoV-19/South Africa/Tygerberg_359_CC2/2020\|EPI_IS | ···················· | ·A·················· | ····················· | ···················· | ···················· | ···················· |
| S | 1763 hCoV-19/South Africa/Tygerberg_365_CC2/2020\|EPI_IS | ···················· | ·A·················· | ····················· | ···················· | ···················· | ···················· |
| S | 1764 hCoV-19/South Africa/Tygerberg_366_CC2/2020\|EPI_IS | ···················· | ···················· | ····················· | ···················· | ···················· | ···················· |
| S | 1765 hCoV-19/South Africa/KRISP-K004275-CE1/2020\|EPI_IS | ···················· | ···················· | ····················· | ···················· | ···················· | ···················· |
| S | 1766 hCoV-19/South Africa/KRISP-K004276-CE1/2020\|EPI_IS | ···················· | ···················· | ····················· | ···················· | ···················· | ···················· |
| S | 1767 hCoV-19/South Africa/KRISP-K004277-CE1/2020\|EPI_IS | ···················· | ···················· | ····················· | ···················· | ···················· | ···················· |
| S | 1768 hCoV-19/South Africa/KRISP-K004278-CE1/2020\|EPI_IS | ···················· | ···················· | ····················· | ···················· | ···················· | ···················· |
| S | 1769 hCoV-19/South Africa/KRISP-K004280-CE1/2020\|EPI_IS | ···················· | ···················· | ····················· | ···················· | ···················· | ···················· |
| S | 1770 hCoV-19/South Africa/KRISP-K004281-CE1/2020\|EPI_IS | ···················· | ···················· | ····················· | ···················· | ···················· | ···················· |
| S | 1771 hCoV-19/South Africa/KRISP-K004282/2020\|EPI_ISL_66 | ···················· | ···················· | ····················· | ···················· | ···················· | ···················· |
| S | 1772 hCoV-19/South Africa/KRISP-K004284/2020\|EPI_ISL_66 | ···················· | ···················· | ····················· | ···················· | ···················· | ···················· |
| S | 1773 hCoV-19/South Africa/KRISP-K004287/2020\|EPI_ISL_66 | ···················· | ···················· | ····················· | ···················· | ···················· | ···················· |
| S | 1774 hCoV-19/South Africa/KRISP-K004288/2020\|EPI_ISL_66 | ···················· | ···················· | ····················· | ···················· | ···················· | ···················· |
| S | 1775 hCoV-19/South Africa/KRISP-K004290/2020\|EPI_ISL_66 | ···················· | ···················· | ····················· | ···················· | ···················· | ···················· |
| S | 1776 hCoV-19/South Africa/KRISP-K004291/2020\|EPI_ISL_66 | ···················· | ···················· | ····················· | ···················· | ···················· | ···················· |
| S | 1777 hCoV-19/South Africa/KRISP-K004292/2020\|EPI_ISL_66 | ···················· | ···················· | ····················· | ···················· | ···················· | ···················· |
| S | 1778 hCoV-19/South Africa/KRISP-K004293/2020\|EPI_ISL_66 | ···················· | ···················· | ····················· | ···················· | ···················· | ···················· |
| S | 1779 hCoV-19/South Africa/KRISP-K004294/2020\|EPI_ISL_66 | ···················· | ···················· | ····················· | ···················· | ···················· | ···················· |
| S | 1780 hCoV-19/South Africa/KRISP-K004295/2020\|EPI_ISL_66 | ···················· | ···················· | ····················· | ···················· | ···················· | ···················· |
| S | 1781 hCoV-19/South Africa/KRISP-K004302/2020\|EPI_ISL_66 | ···················· | ···················· | ····················· | ···················· | ···················· | ···················· |
| S | 1783 hCoV-19/South Africa/KRISP-K004311/2020\|EPI_ISL_66 | ···················· | ···················· | ····················· | ···················· | ···················· | ···················· |
| S | 1784 hCoV-19/South Africa/KRISP-K004313/2020\|EPI_ISL_66 | ···················· | ···················· | ····················· | ···················· | ···················· | ···················· |
| S | 1785 hCoV-19/South Africa/KRISP-K004315/2020\|EPI_ISL_66 | ···················· | ···················· | ····················· | ···················· | ···················· | ···················· |
| S | 1786 hCoV-19/South Africa/KRISP-K004316/2020\|EPI_ISL_66 | ···················· | ·A·················· | ····················· | ···················· | ···················· | ···················· |
| S | 1787 hCoV-19/South Africa/KRISP-K004317/2020\|EPI_ISL_66 | ···················· | ·A·················· | ····················· | ···················· | ···················· | ···················· |
| S | 1788 hCoV-19/South Africa/KRISP-K004318/2020\|EPI_ISL_66 | ···················· | ·A·················· | ····················· | ···················· | ···················· | ···················· |
| S | 1789 hCoV-19/South Africa/KRISP-K004320/2020\|EPI_ISL_66 | ···················· | ···················· | ····················· | ···················· | ···················· | ···················· |
| S | 1790 hCoV-19/South Africa/KRISP-K004321/2020\|EPI_ISL_66 | ···················· | ···················· | ····················· | ···················· | ···················· | ···················· |
| S | 1791 hCoV-19/South Africa/KRISP-K004322/2020\|EPI_ISL_66 | ···················· | ···················· | ····················· | ···················· | ···················· | ···················· |
| S | 1792 hCoV-19/South Africa/KRISP-K004323/2020\|EPI_ISL_66 | ···················· | ·A·················· | ····················· | ···················· | ···················· | ···················· |
| S | 1793 hCoV-19/Scotland/QEUH-AD543B/2020\|EPI_ISL_649701\|2 | ···················· | ···················· | ····················· | ···················· | ···················· | ···················· |
| S | 1794 hCoV-19/Scotland/QEUH-AD145D/2020\|EPI_ISL_649702\|2 | ···················· | ···················· | ····················· | ···················· | ···················· | ···················· |
| S | 1795 hCoV-19/Scotland/QEUH-AD429C/2020\|EPI_ISL_649703\|2 | ···················· | ···················· | ····················· | ···················· | ···················· | ···················· |
| S | 1796 hCoV-19/Scotland/QEUH-AD5459/2020\|EPI_ISL_649704\|2 | ···················· | ···················· | ····················· | ···················· | ···················· | ···················· |
| S | 1797 hCoV-19/Scotland/QEUH-AD56F9/2020\|EPI_ISL_649705\|2 | ···················· | ···················· | ····················· | ···················· | ···················· | ···················· |
| S | 1798 hCoV-19/Scotland/QEUH-AD5A2A/2020\|EPI_ISL_649706\|2 | ···················· | ···················· | ····················· | ···················· | ···················· | ···················· |
| S | 1799 hCoV-19/Scotland/QEUH-AD1727/2020\|EPI_ISL_649707\|2 | ···················· | ···················· | ····················· | ···················· | ···················· | ···················· |

```
S 1800 hCoV-19/Scotland/QEUH-AD195E/2020|EPI_ISL_649708|2 .................... .................... .................... .................... .................... ....................
S 1801 hCoV-19/Scotland/QEUH-AD137E/2020|EPI_ISL_649709|2 .................... .................... .................... .................... .................... ....................
S 1802 hCoV-19/Scotland/QEUH-AD1AC4/2020|EPI_ISL_649711|2 .................... .................... .................... .................... .................... ....................
S 1803 hCoV-19/Scotland/QEUH-AD13BA/2020|EPI_ISL_649713|2 .................... .................... .................... .................... .................... ....................
S 1804 hCoV-19/Scotland/QEUH-AD1305/2020|EPI_ISL_649714|2 .................... .................... .................... .................... .................... ....................
S 1805 hCoV-19/Scotland/QEUH-AD1402/2020|EPI_ISL_649717|2 .................... .................... .................... .................... .................... ....................
S 1806 hCoV-19/Scotland/QEUH-AD13AB/2020|EPI_ISL_649718|2 .................... .................... .................... .................... .................... ....................
S 1807 hCoV-19/Scotland/QEUH-AD088F/2020|EPI_ISL_649719|2 .................... .................... .................... .................... .................... ....................
S 1808 hCoV-19/Scotland/QEUH-AD13E7/2020|EPI_ISL_649720|2 .................... .................... .................... .................... .................... ....................
S 1809 hCoV-19/Scotland/QEUH-AD136F/2020|EPI_ISL_649721|2 .................... .................... .................... .................... .................... ....................
S 1810 hCoV-19/Scotland/QEUH-AD139C/2020|EPI_ISL_649723|2 .................... .................... .................... .................... .................... ....................
S 1811 hCoV-19/Scotland/QEUH-AD1596/2020|EPI_ISL_649724|2 .................... .................... .................... .................... .................... ....................
S 1812 hCoV-19/Scotland/QEUH-AD6652/2020|EPI_ISL_649727|2 .................... .................... .................... .................... .................... ....................
S 1813 hCoV-19/Spain/MD-IBV-99008625/2020|EPI_ISL_654200| .................... .................... .................... .................... .................... ....................
S 1814 hCoV-19/Spain/MD-IBV-99008627/2020|EPI_ISL_654201| .................... .................... .................... .................... .................... ....................
S 1816 hCoV-19/Spain/MD-IBV-99008629/2020|EPI_ISL_654203| .................... .................... .................... .................... .................... ....................
S 1817 hCoV-19/Spain/MD-IBV-99008630/2020|EPI_ISL_654204| .................... .................... .................... .................... .................... ....................
S 1818 hCoV-19/Spain/MD-IBV-99008633/2020|EPI_ISL_654205| .................... .................... .................... .................... ......T............. ....................
S 1820 hCoV-19/Spain/MD-IBV-99008636/2020|EPI_ISL_654207| .................... .................... .................... .................... .................... ....................
S 1828 hCoV-19/Spain/MD-IBV-99008651/2020|EPI_ISL_654215| .................... .................... .................... .................... .................... ....................
S 1829 hCoV-19/Spain/MD-IBV-99008652/2020|EPI_ISL_654216| .................... .................... .................... .................... .................... ....................
S 1830 hCoV-19/Spain/MD-IBV-99008654/2020|EPI_ISL_654217| .................... .................... .................... .................... .................... ....................
S 1831 hCoV-19/Spain/MD-IBV-99008656/2020|EPI_ISL_654218| .................... .................... .................... .................... .................... ....................
S 1832 hCoV-19/Spain/MD-IBV-99008657/2020|EPI_ISL_654219| .................... .................... .................... .................... .................... ....................
S 1834 hCoV-19/Spain/MD-IBV-99008663/2020|EPI_ISL_654224| .................... .................... .................... .................... .................... ....................
S 1835 hCoV-19/Spain/MD-IBV-99008666/2020|EPI_ISL_654227| .................... .................... .................... .................... .................... ....................
S 1838 hCoV-19/Spain/MD-IBV-99008671/2020|EPI_ISL_654230| .................... .................... .................... .................... .................... ....................
S 1839 hCoV-19/Spain/MD-IBV-99008676/2020|EPI_ISL_654231| .................... .................... .................... .................... .................... ....................
S 1842 hCoV-19/Spain/MD-IBV-99008679/2020|EPI_ISL_654234| .................... .................... .................... .................... .................... ....................
S 1848 hCoV-19/Spain/MD-IBV-99008685/2020|EPI_ISL_654240| .................... .................... .................... .................... .................... ....................
S 1849 hCoV-19/Spain/MD-IBV-99008686/2020|EPI_ISL_654241| .................... .................... .................... .................... .................... ....................
S 1858 hCoV-19/Spain/MD-IBV-99008699/2020|EPI_ISL_654279| .................... .................... .................... .................... .................... ....................
S 1859 hCoV-19/Spain/MD-IBV-99008700/2020|EPI_ISL_654280| .................... .................... .................... .................... .................... ....................
S 1860 hCoV-19/Spain/MD-IBV-99008701/2020|EPI_ISL_654281| .................... .................... .................... .................... .................... ....................
S 1861 hCoV-19/Spain/MD-IBV-99008702/2020|EPI_ISL_654282| .................... .................... .................... .................... .................... ....................
S 1863 hCoV-19/Spain/MD-IBV-99008704/2020|EPI_ISL_654284| .................... .................... .................... .................... .................... ....................
S 1864 hCoV-19/Spain/MD-IBV-99008707/2020|EPI_ISL_654285| .................... .................... .................... .................... .................... ....................
S 1865 hCoV-19/Spain/MD-IBV-99008708/2020|EPI_ISL_654286| .................... .................... .................... .................... .................... ....................
S 1867 hCoV-19/Spain/MD-IBV-99008710/2020|EPI_ISL_654288| .................... .................... .................... .................... .................... ....................
S 1868 hCoV-19/Spain/MD-IBV-99008711/2020|EPI_ISL_654289| .................... .................... .................... .................... .................... ....................
S 1869 hCoV-19/Spain/MD-IBV-99008712/2020|EPI_ISL_654290| .................... .................... .................... .................... .................... ....................
S 1870 hCoV-19/Spain/MD-IBV-99008714/2020|EPI_ISL_654291| .................... .................... .................... .................... .................... ....................
S 1871 hCoV-19/Spain/MD-IBV-99008715/2020|EPI_ISL_654292| .................... .................... .................... .................... .................... ....................
S 1875 hCoV-19/Spain/MD-IBV-99008719/2020|EPI_ISL_654296| .................... .................... .................... .................... .................... ....................
S 1877 hCoV-19/Spain/MD-IBV-99008723/2020|EPI_ISL_654298| .................... .................... .................... .................... .................... ....................
S 1879 hCoV-19/England/ALDP-B5B496/2020|EPI_ISL_655501|20 .................... .................... .................... .................... .................... ....................
S 1880 hCoV-19/England/ALDP-B63D81/2020|EPI_ISL_655502|20 .................... .................... .................... .................... .................... ....................
S 1881 hCoV-19/England/ALDP-B62EF8/2020|EPI_ISL_655503|20 .................... .................... .................... .................... .................... ....................
S 1882 hCoV-19/England/ALDP-B5B96A/2020|EPI_ISL_655505|20 .................... .................... .................... .................... .................... ....................
S 1883 hCoV-19/England/ALDP-B62DBF/2020|EPI_ISL_655506|20 .................... .................... .................... .................... .................... ....................
S 1884 hCoV-19/England/ALDP-B5E721/2020|EPI_ISL_655507|20 .................... .................... .................... .................... .................... ....................
S 1885 hCoV-19/England/ALDP-B62DEC/2020|EPI_ISL_655508|20 .................... .................... .................... .................... .................... ....................
S 1886 hCoV-19/England/ALDP-B5BA1C/2020|EPI_ISL_655509|20 .................... .................... .................... .................... .................... ....................
S 1887 hCoV-19/England/ALDP-B5B3D5/2020|EPI_ISL_655510|20 .................... .................... .................... .................... .................... ....................
S 1888 hCoV-19/England/ALDP-B5AF5A/2020|EPI_ISL_655512|20 .................... .................... .................... .................... .................... ....................
S 1889 hCoV-19/England/ALDP-B62F40/2020|EPI_ISL_655514|20 .................... .................... .................... .................... .................... ....................
S 1890 hCoV-19/England/ALDP-B62EAD/2020|EPI_ISL_655515|20 .................... .................... .................... .................... .................... ....................
S 1891 hCoV-19/England/ALDP-B5B302/2020|EPI_ISL_655516|20 .................... .................... .................... .................... .................... ....................
S 1892 hCoV-19/England/ALDP-B5B19F/2020|EPI_ISL_655517|20 .................... .A.................. .................... .................... .................... ...A................
S 1893 hCoV-19/England/ALDP-B5B645/2020|EPI_ISL_655519|20 .................... .................... .................... .................... .................... ....................
S 1894 hCoV-19/England/ALDP-B5E712/2020|EPI_ISL_655520|20 .................... .................... .................... .................... .................... ....................
S 1895 hCoV-19/England/ALDP-B62F22/2020|EPI_ISL_655522|20 .................... .................... .................... .................... .................... ....................
S 1896 hCoV-19/England/ALDP-B5B733/2020|EPI_ISL_655524|20 .................... .................... .................... .................... .................... ....................
S 1897 hCoV-19/England/ALDP-B5B9C4/2020|EPI_ISL_655526|20 .................... .................... .................... .................... .................... ....................
S 1898 hCoV-19/England/ALDP-B5B2AB/2020|EPI_ISL_655527|20 .................... .................... .................... .................... .................... ....................
S 1899 hCoV-19/England/ALDP-B63D09/2020|EPI_ISL_655528|20 .................... .................... .................... .................... .................... ....................
S 1900 hCoV-19/England/ALDP-B62CFE/2020|EPI_ISL_655529|20 .................... .................... .................... .................... ......T............. ....................
S 1901 hCoV-19/England/ALDP-B5B672/2020|EPI_ISL_655530|20 .................... .................... .................... .................... .................... ....................
S 1902 hCoV-19/England/ALDP-B5AFB4/2020|EPI_ISL_655531|20 .................... .................... .................... .................... .................... ....................
S 1903 hCoV-19/England/ALDP-B5B074/2020|EPI_ISL_655532|20 .................... .................... .................... .................... .................... ....................
S 1904 hCoV-19/England/ALDP-B5B5DF/2020|EPI_ISL_655533|20 .................... .................... .................... .................... .................... ....................
S 1905 hCoV-19/England/ALDP-B62AB8/2020|EPI_ISL_655534|20 .................... .................... .................... .................... .................... ....................
S 1906 hCoV-19/England/ALDP-B5B803/2020|EPI_ISL_655535|20 .................... .................... .................... .................... .................... ....................
S 1907 hCoV-19/England/ALDP-B5BB19/2020|EPI_ISL_655536|20 .................... .................... .................... .................... .................... ....................
S 1908 hCoV-19/England/ALDP-B62E34/2020|EPI_ISL_655538|20 .................... .................... .................... .................... .................... ....................
S 1909 hCoV-19/England/ALDP-B5B083/2020|EPI_ISL_655540|20 .................... .................... .................... .................... .................... ....................
S 1910 hCoV-19/England/ALDP-B5B9D3/2020|EPI_ISL_655541|20 .................... .................... .................... .................... .................... ....................
S 1911 hCoV-19/England/ALDP-B5AF87/2020|EPI_ISL_655542|20 .................... .................... .................... .................... .................... ....................
S 1912 hCoV-19/England/ALDP-B5AF96/2020|EPI_ISL_655543|20 .................... .................... .................... .................... .................... ....................
S 1913 hCoV-19/England/ALDP-B5AEF3/2020|EPI_ISL_655545|20 .................... .................... .................... .................... .................... ....................
```

| | | | | | | | |
|---|---|---|---|---|---|---|---|
| S | 1914 hCoV-19/England/ALDP-B62DA0/2020\|EPI_ISL_655546\|20 | ···················· | ···················· | ···················· | ···················· | ···················· | ···················· |
| S | 1915 hCoV-19/England/ALDP-B5AFA5/2020\|EPI_ISL_655547\|20 | ···················· | ···················· | ···················· | ···················· | ·····T·············· | ···················· |
| S | 1916 hCoV-19/England/ALDP-B5B1AE/2020\|EPI_ISL_655548\|20 | ···················· | ···················· | ···················· | ···················· | ···················· | ···················· |
| S | 1917 hCoV-19/England/ALDP-B5BB0A/2020\|EPI_ISL_655549\|20 | ···················· | ···················· | ···················· | ···················· | ···················· | ···················· |
| S | 1918 hCoV-19/England/ALDP-B62BF1/2020\|EPI_ISL_655554\|20 | ···················· | ···················· | ···················· | ···················· | ···················· | ···················· |
| S | 1919 hCoV-19/England/ALDP-B5B2D8/2020\|EPI_ISL_655558\|20 | ···················· | ···················· | ···················· | ···················· | ···················· | ···················· |
| S | 1920 hCoV-19/England/ALDP-B5B01A/2020\|EPI_ISL_655559\|20 | ···················· | ···················· | ···················· | ···················· | ···················· | ···················· |
| S | 1921 hCoV-19/England/ALDP-B5B812/2020\|EPI_ISL_655561\|20 | ···················· | ···················· | ···················· | ···················· | ···················· | ···················· |
| S | 1922 hCoV-19/England/ALDP-B5BCAD/2020\|EPI_ISL_655562\|20 | ···················· | ···················· | ···················· | ···················· | ···················· | ···················· |
| S | 1923 hCoV-19/England/ALDP-B5B8D6/2020\|EPI_ISL_655563\|20 | ···················· | ···················· | ···················· | ···················· | ···················· | ···················· |
| S | 1924 hCoV-19/England/ALDP-B5B37B/2020\|EPI_ISL_655564\|20 | ···················· | ···················· | ···················· | ···················· | ···················· | ···················· |
| S | 1925 hCoV-19/England/ALDP-B62BB5/2020\|EPI_ISL_655565\|20 | ···················· | ···················· | ···················· | ···················· | ···················· | ···················· |
| S | 1926 hCoV-19/England/ALDP-B5B241/2020\|EPI_ISL_655566\|20 | ···················· | ···················· | ···················· | ···················· | ···················· | ···················· |
| S | 1927 hCoV-19/England/ALDP-B5B1F9/2020\|EPI_ISL_655567\|20 | ···················· | ···················· | ···················· | ···················· | ···················· | ···················· |
| S | 1928 hCoV-19/England/ALDP-B5B0FC/2020\|EPI_ISL_655568\|20 | ···················· | ···················· | ···················· | ···················· | ···················· | ···················· |
| S | 1929 hCoV-19/England/ALDP-B5B89A/2020\|EPI_ISL_655569\|20 | ···················· | ···················· | ···················· | ···················· | ···················· | ···················· |
| S | 1930 hCoV-19/England/ALDP-B5B092/2020\|EPI_ISL_655570\|20 | ···················· | ···················· | ···················· | ···················· | ···················· | ···················· |
| S | 1931 hCoV-19/England/ALDP-B62C67/2020\|EPI_ISL_655571\|20 | ···················· | ···················· | ···················· | ···················· | ···················· | ···················· |
| S | 1932 hCoV-19/England/ALDP-B6305B/2020\|EPI_ISL_655572\|20 | ···················· | ···················· | ···················· | ···················· | ···················· | ···················· |
| S | 1933 hCoV-19/England/ALDP-B5B2C9/2020\|EPI_ISL_655573\|20 | ···················· | ···················· | ···················· | ···················· | ···················· | ···················· |
| S | 1934 hCoV-19/England/ALDP-B5BAFE/2020\|EPI_ISL_655574\|20 | ···················· | ···················· | ···················· | ···················· | ···················· | ···················· |
| S | 1935 hCoV-19/England/ALDP-B5B87C/2020\|EPI_ISL_655575\|20 | ···················· | ···················· | ···················· | ···················· | ···················· | ···················· |
| S | 1936 hCoV-19/England/ALDP-B63D63/2020\|EPI_ISL_655576\|20 | ···················· | ···················· | ···················· | ···················· | ···················· | ···················· |
| S | 1937 hCoV-19/England/ALDP-B5B9E2/2020\|EPI_ISL_655577\|20 | ···················· | ···················· | ···················· | ···················· | ···················· | ···················· |
| S | 1938 hCoV-19/England/ALDP-B62ECB/2020\|EPI_ISL_655578\|20 | ···················· | ·A·················· | ···················· | ···················· | ···················· | ···················· |
| S | 1939 hCoV-19/England/ALDP-B62BA6/2020\|EPI_ISL_655580\|20 | ···················· | ···················· | ···················· | ···················· | ···················· | ···················· |
| S | 1940 hCoV-19/England/ALDP-B5B50C/2020\|EPI_ISL_655583\|20 | ···················· | ···················· | ···················· | ···················· | ···················· | ···················· |
| S | 1941 hCoV-19/England/ALDP-B5B9B5/2020\|EPI_ISL_655584\|20 | ···················· | ···················· | ···················· | ···················· | ···················· | ···················· |
| S | 1942 hCoV-19/England/ALDP-B63D45/2020\|EPI_ISL_655585\|20 | ···················· | ···················· | ···················· | ···················· | ···················· | ···················· |
| S | 1943 hCoV-19/England/ALDP-B5B153/2020\|EPI_ISL_655589\|20 | ···················· | ···················· | ···················· | ···················· | ···················· | ···················· |
| S | 1944 hCoV-19/England/ALDP-B5B3E4/2020\|EPI_ISL_655590\|20 | ···················· | ···················· | ···················· | ···················· | ···················· | ···················· |
| S | 1945 hCoV-19/England/ALDP-B5B6FA/2020\|EPI_ISL_655591\|20 | ···················· | ···················· | ···················· | ···················· | ···················· | ···················· |
| S | 1946 hCoV-19/England/ALDP-B5B5C0/2020\|EPI_ISL_655592\|20 | ···················· | ···················· | ···················· | ···················· | ···················· | ···················· |
| S | 1947 hCoV-19/England/ALDP-B5B566/2020\|EPI_ISL_655593\|20 | ···················· | ···················· | ···················· | ···················· | ···················· | ···················· |
| S | 1948 hCoV-19/England/ALDP-B5BC43/2020\|EPI_ISL_655594\|20 | ···················· | ···················· | ···················· | ···················· | ···················· | ···················· |
| S | 1949 hCoV-19/England/ALDP-B5BC16/2020\|EPI_ISL_655595\|20 | ···················· | ···················· | ···················· | ···················· | ···················· | ···················· |
| S | 1950 hCoV-19/England/ALDP-B62C3A/2020\|EPI_ISL_655597\|20 | ···················· | ···················· | ···················· | ···················· | ···················· | ···················· |
| S | 1951 hCoV-19/England/ALDP-B5B690/2020\|EPI_ISL_655598\|20 | ···················· | ···················· | ···················· | ···················· | ···················· | ···················· |
| S | 1952 hCoV-19/England/ALDP-B62B4C/2020\|EPI_ISL_655599\|20 | ···················· | ···················· | ···················· | ···················· | ···················· | ···················· |
| S | 1953 hCoV-19/England/CAMC-B5366B/2020\|EPI_ISL_656812\|20 | ···················· | ···················· | ···················· | ···················· | ···················· | ···················· |
| S | 1954 hCoV-19/England/CAMC-B5403F/2020\|EPI_ISL_656813\|20 | ···················· | ···················· | ···················· | ···················· | ···················· | ···················· |
| S | 1955 hCoV-19/England/CAMC-B538FC/2020\|EPI_ISL_656814\|20 | ···················· | ···················· | ···················· | ···················· | ·····T·············· | ···················· |
| S | 1956 hCoV-19/England/CAMC-B534BC/2020\|EPI_ISL_656816\|20 | ···················· | ···················· | ···················· | ···················· | ···················· | ···················· |
| S | 1957 hCoV-19/England/CAMC-B538CF/2020\|EPI_ISL_656817\|20 | ···················· | ···················· | ···················· | ···················· | ···················· | ···················· |
| S | 1958 hCoV-19/England/CAMC-B52356/2020\|EPI_ISL_656818\|20 | ···················· | ···················· | ···················· | ···················· | ···················· | ···················· |
| S | 1959 hCoV-19/England/CAMC-B53874/2020\|EPI_ISL_656820\|20 | ···················· | ···················· | ···················· | ···················· | ···················· | ···················· |
| S | 1960 hCoV-19/England/CAMC-B53838/2020\|EPI_ISL_656821\|20 | ···················· | ···················· | ···················· | ···················· | ···················· | ···················· |
| S | 1961 hCoV-19/England/CAMC-B5448E/2020\|EPI_ISL_656822\|20 | ···················· | ···················· | ···················· | ···················· | ···················· | ···················· |
| S | 1962 hCoV-19/England/MILK-B56CC0/2020\|EPI_ISL_656824\|20 | ···················· | ···················· | ···················· | ···················· | ···················· | ···················· |
| S | 1963 hCoV-19/England/CAMC-B522D1/2020\|EPI_ISL_656825\|20 | ···················· | ···················· | ···················· | ···················· | ···················· | ···················· |
| S | 1964 hCoV-19/England/CAMC-B5362F/2020\|EPI_ISL_656826\|20 | ···················· | ···················· | ···················· | ···················· | ···················· | ···················· |
| S | 1965 hCoV-19/England/MILK-B56C75/2020\|EPI_ISL_656827\|20 | ···················· | ···················· | ···················· | ···················· | ···················· | ···················· |
| S | 1966 hCoV-19/England/CAMC-B542B1/2020\|EPI_ISL_656828\|20 | ···················· | ·A·················· | ···················· | ···················· | ···················· | ···················· |
| S | 1967 hCoV-19/England/MILK-B569AB/2020\|EPI_ISL_656829\|20 | ···················· | ···················· | ···················· | ···················· | ·····T·············· | ···················· |
| S | 1968 hCoV-19/England/CAMC-B53768/2020\|EPI_ISL_656830\|20 | ···················· | ···················· | ···················· | ···················· | ···················· | ···················· |
| S | 1969 hCoV-19/England/CAMC-B536F2/2020\|EPI_ISL_656833\|20 | ···················· | ···················· | ···················· | ···················· | ···················· | ···················· |
| S | 1970 hCoV-19/England/MILK-B56AA8/2020\|EPI_ISL_656834\|20 | ···················· | ···················· | ···················· | ···················· | ···················· | ···················· |
| S | 1971 hCoV-19/England/MILK-B56C57/2020\|EPI_ISL_656835\|20 | ···················· | ···················· | ···················· | ···················· | ···················· | ···················· |
| S | 1972 hCoV-19/England/CAMC-B52778/2020\|EPI_ISL_656836\|20 | ···················· | ···················· | ···················· | ···················· | ···················· | ···················· |
| S | 1973 hCoV-19/England/CAMC-B536A7/2020\|EPI_ISL_656837\|20 | ···················· | ···················· | ···················· | ···················· | ···················· | ···················· |
| S | 1974 hCoV-19/England/MILK-B56C2A/2020\|EPI_ISL_656838\|20 | ···················· | ···················· | ···················· | ···················· | ···················· | ···················· |
| S | 1975 hCoV-19/England/CAMC-B53FE8/2020\|EPI_ISL_656839\|20 | ···················· | ···················· | ···················· | ···················· | ···················· | ···················· |
| S | 1976 hCoV-19/England/MILK-B56835/2020\|EPI_ISL_656840\|20 | ···················· | ···················· | ···················· | ···················· | ···················· | ···················· |
| S | 1977 hCoV-19/England/MILK-B56D72/2020\|EPI_ISL_656841\|20 | ···················· | ···················· | ···················· | ···················· | ···················· | ···················· |
| S | 1978 hCoV-19/England/CAMC-B525D8/2020\|EPI_ISL_656842\|20 | ···················· | ···················· | ···················· | ···················· | ···················· | ···················· |
| S | 1979 hCoV-19/England/CAMC-B52383/2020\|EPI_ISL_656843\|20 | ···················· | ···················· | ···················· | ···················· | ···················· | ···················· |
| S | 1980 hCoV-19/England/MILK-B56CA2/2020\|EPI_ISL_656846\|20 | ···················· | ···················· | ···················· | ···················· | ···················· | ···················· |
| S | 1981 hCoV-19/England/CAMC-B5422A/2020\|EPI_ISL_656847\|20 | ···················· | ···················· | ···················· | ···················· | ···················· | ···················· |
| S | 1982 hCoV-19/England/MILK-B56880/2020\|EPI_ISL_656848\|20 | ···················· | ···················· | ···················· | ···················· | ···················· | ···················· |
| S | 1983 hCoV-19/England/CAMC-B526D5/2020\|EPI_ISL_656850\|20 | ···················· | ···················· | ···················· | ···················· | ···················· | ···················· |
| S | 1984 hCoV-19/England/CAMC-B54187/2020\|EPI_ISL_656851\|20 | ···················· | ···················· | ···················· | ···················· | ···················· | ···················· |
| S | 1985 hCoV-19/England/CAMC-B544F7/2020\|EPI_ISL_656852\|20 | ···················· | ···················· | ···················· | ···················· | ···················· | ···················· |
| S | 1986 hCoV-19/England/CAMC-B53883/2020\|EPI_ISL_656853\|20 | ···················· | ···················· | ···················· | ···················· | ·····T·············· | ···················· |
| S | 1987 hCoV-19/England/CAMC-B5259C/2020\|EPI_ISL_656854\|20 | ···················· | ···················· | ···················· | ···················· | ···················· | ···················· |
| S | 1988 hCoV-19/England/CAMC-B536B6/2020\|EPI_ISL_656856\|20 | ···················· | ···················· | ···················· | ···················· | ···················· | ···················· |
| S | 1989 hCoV-19/England/CAMC-B522B3/2020\|EPI_ISL_656857\|20 | ···················· | ···················· | ···················· | ···················· | ···················· | ···················· |
| S | 1990 hCoV-19/England/CAMC-B54196/2020\|EPI_ISL_656858\|20 | ···················· | ···················· | ···················· | ···················· | ···················· | ···················· |
| S | 1991 hCoV-19/England/CAMC-B522A4/2020\|EPI_ISL_656859\|20 | ···················· | ···················· | ···················· | ···················· | ···················· | ···················· |
| S | 1992 hCoV-19/England/MILK-B56BB4/2020\|EPI_ISL_656860\|20 | ···················· | ···················· | ···················· | ···················· | ···················· | ···················· |
| S | 1993 hCoV-19/England/MILK-B56747/2020\|EPI_ISL_656862\|20 | ···················· | ···················· | ···················· | ···················· | ···················· | ···················· |

| | | | | | | | |
|---|---|---|---|---|---|---|---|
| S | 1994 hCoV-19/England/CAMC-B52699/2020\|EPI_ISL_656863\|20 | ···················· | ···················· | ···················· | ···················· | ···················· | ···················· |
| S | 1995 hCoV-19/England/CAMC-B53540/2020\|EPI_ISL_656864\|20 | ···················· | ···················· | ···················· | ···················· | ·····T·············· | ···················· |
| S | 1996 hCoV-19/England/MILK-B56A7B/2020\|EPI_ISL_656865\|20 | ···················· | ···················· | ···················· | ···················· | ···················· | ···················· |
| S | 1997 hCoV-19/England/CAMC-B5268A/2020\|EPI_ISL_656866\|20 | ···················· | ···················· | ···················· | ···················· | ···················· | ···················· |
| S | 1998 hCoV-19/England/MILK-B56A11/2020\|EPI_ISL_656867\|20 | ···················· | ···················· | ···················· | ···················· | ···················· | ···················· |
| S | 1999 hCoV-19/England/CAMC-B5421B/2020\|EPI_ISL_656868\|20 | ···················· | ···················· | ···················· | ···················· | ···················· | ···················· |
| S | 2000 hCoV-19/England/MILK-B56D90/2020\|EPI_ISL_656869\|20 | ···················· | ···················· | ···················· | ···················· | ···················· | ···················· |
| S | 2001 hCoV-19/England/MILK-B56B2D/2020\|EPI_ISL_656871\|20 | ···················· | ···················· | ···················· | ···················· | ···················· | ···················· |
| S | 2002 hCoV-19/England/CAMC-B53382/2020\|EPI_ISL_656872\|20 | ···················· | ···················· | ···················· | ···················· | ·····T·············· | ···················· |
| S | 2003 hCoV-19/England/CAMC-B5363E/2020\|EPI_ISL_656873\|20 | ···················· | ···················· | ···················· | ···················· | ···················· | ···················· |
| S | 2004 hCoV-19/England/CAMC-B54336/2020\|EPI_ISL_656874\|20 | ···················· | ···················· | ···················· | ···················· | ···················· | ···················· |
| S | 2005 hCoV-19/England/CAMC-B54266/2020\|EPI_ISL_656875\|20 | ···················· | ···················· | ···················· | ···················· | ···················· | ···················· |
| S | 2006 hCoV-19/England/CAMC-B5263F/2020\|EPI_ISL_656876\|20 | ···················· | ···················· | ···················· | ···················· | ···················· | ···················· |
| S | 2007 hCoV-19/England/MILK-B56CB1/2020\|EPI_ISL_656877\|20 | ···················· | ···················· | ···················· | ···················· | ···················· | ···················· |
| S | 2008 hCoV-19/England/CAMC-B540B7/2020\|EPI_ISL_656878\|20 | ···················· | ···················· | ···················· | ···················· | ···················· | ···················· |
| S | 2009 hCoV-19/England/MILK-B56D45/2020\|EPI_ISL_656879\|20 | ···················· | ···················· | ···················· | ···················· | ···················· | ···················· |
| S | 2010 hCoV-19/England/CAMC-B5231A/2020\|EPI_ISL_656889\|20 | ···················· | ···················· | ···················· | ···················· | ···················· | ···················· |
| S | 2011 hCoV-19/England/CAMC-B524BD/2020\|EPI_ISL_656890\|20 | ···················· | ···················· | ···················· | ···················· | ···················· | ···················· |
| S | 2012 hCoV-19/England/CAMC-B53917/2020\|EPI_ISL_656891\|20 | ···················· | ···················· | ···················· | ···················· | ···················· | ···················· |
| S | 2013 hCoV-19/England/MILK-B56AD5/2020\|EPI_ISL_656892\|20 | ···················· | ···················· | ···················· | ···················· | ···················· | ···················· |
| S | 2014 hCoV-19/England/MILK-B56D09/2020\|EPI_ISL_656893\|20 | ···················· | ···················· | ···················· | ···················· | ···················· | ···················· |
| S | 2015 hCoV-19/England/CAMC-B524DB/2020\|EPI_ISL_656896\|20 | ···················· | ···················· | ···················· | ···················· | ···················· | ···················· |
| S | 2016 hCoV-19/England/CAMC-B534F8/2020\|EPI_ISL_656898\|20 | ···················· | ···················· | ···················· | ···················· | ···················· | ···················· |
| S | 2017 hCoV-19/England/CAMC-B541C3/2020\|EPI_ISL_656899\|20 | ···················· | ···················· | ···················· | ···················· | ···················· | ···················· |
| S | 2018 hCoV-19/England/CAMC-B37D95/2020\|EPI_ISL_659090\|20 | ···················· | ···················· | ···················· | ···················· | ···················· | ···················· |
| S | 2019 hCoV-19/England/CAMC-B37F71/2020\|EPI_ISL_659091\|20 | ···················· | ···················· | ···················· | ···················· | ···················· | ···················· |
| S | 2020 hCoV-19/England/MILK-B38E46/2020\|EPI_ISL_659092\|20 | ···················· | ···················· | ···················· | ···················· | ···················· | ···················· |
| S | 2021 hCoV-19/England/CAMC-B37FDB/2020\|EPI_ISL_659093\|20 | ···················· | ···················· | ···················· | ···················· | ···················· | ···················· |
| S | 2022 hCoV-19/England/CAMC-B38013/2020\|EPI_ISL_659094\|20 | ···················· | ···················· | ···················· | ···················· | ···················· | ···················· |
| S | 2023 hCoV-19/England/MILK-B391C4/2020\|EPI_ISL_659095\|20 | ···················· | ···················· | ···················· | ···················· | ···················· | ···················· |
| S | 2024 hCoV-19/England/MILK-B3965C/2020\|EPI_ISL_659096\|20 | ···················· | ···················· | ···················· | ···················· | ···················· | ···················· |
| S | 2025 hCoV-19/England/CAMC-B33810/2020\|EPI_ISL_659097\|20 | ···················· | ···················· | ···················· | ···················· | ···················· | ···················· |
| S | 2026 hCoV-19/England/CAMC-B32477/2020\|EPI_ISL_659098\|20 | ···················· | ···················· | ···················· | ···················· | ···················· | ···················· |
| S | 2027 hCoV-19/England/CAMC-B339A4/2020\|EPI_ISL_659099\|20 | ···················· | ···················· | ···················· | ···················· | ·····T·············· | ···················· |
| S | 2028 hCoV-19/USA/MD-MDH-0381/2020\|EPI_ISL_660079\|2020-1 | ···················· | ·A·················· | ···················· | ···················· | ···················· | ···················· |
| S | 2029 hCoV-19/USA/MD-MDH-0382/2020\|EPI_ISL_660080\|2020-1 | ···················· | ·A·················· | ···················· | ···················· | ···················· | ···················· |
| S | 2030 hCoV-19/USA/MD-MDH-0384/2020\|EPI_ISL_660081\|2020-1 | ···················· | ·A·················· | ···················· | ···················· | ···················· | ···················· |
| S | 2031 hCoV-19/USA/MD-MDH-0385/2020\|EPI_ISL_660082\|2020-1 | ···················· | ···················· | ···················· | ···················· | ···················· | ···················· |
| S | 2032 hCoV-19/USA/MD-MDH-0388/2020\|EPI_ISL_660083\|2020-1 | ···················· | ···················· | ···················· | ···················· | ···················· | ···················· |
| S | 2033 hCoV-19/USA/MD-MDH-0389/2020\|EPI_ISL_660084\|2020-1 | ···················· | ···················· | ···················· | ···················· | ···················· | ···················· |
| S | 2034 hCoV-19/USA/MD-MDH-0391/2020\|EPI_ISL_660085\|2020-1 | ···················· | ···················· | ···················· | ···················· | ···················· | ···················· |
| S | 2035 hCoV-19/USA/MD-MDH-0392/2020\|EPI_ISL_660086\|2020-1 | ···················· | ············C······· | ···················· | ···················· | ···················· | ···················· |
| S | 2036 hCoV-19/USA/MD-MDH-0393/2020\|EPI_ISL_660087\|2020-1 | ···················· | ············C······· | ···················· | ···················· | ···················· | ···················· |
| S | 2037 hCoV-19/USA/MD-MDH-0394/2020\|EPI_ISL_660088\|2020-1 | ···················· | ·A·················· | ···················· | ···················· | ···················· | ···················· |
| S | 2038 hCoV-19/USA/MD-MDH-0395/2020\|EPI_ISL_660089\|2020-1 | ···················· | ············C······· | ···················· | ···················· | ···················· | ···················· |
| S | 2039 hCoV-19/USA/MD-MDH-0396/2020\|EPI_ISL_660090\|2020-1 | ···················· | ············C······· | ···················· | ···················· | ···················· | ···················· |
| S | 2040 hCoV-19/USA/MD-MDH-0397/2020\|EPI_ISL_660091\|2020-1 | ···················· | ···················· | ···················· | ···················· | ···················· | ···················· |
| S | 2041 hCoV-19/USA/MD-MDH-0398/2020\|EPI_ISL_660092\|2020-1 | ···················· | ···················· | ···················· | ···················· | ···················· | ···················· |
| S | 2042 hCoV-19/USA/MD-MDH-0399/2020\|EPI_ISL_660093\|2020-1 | ···················· | ···················· | ···················· | ···················· | ···················· | ···················· |
| S | 2043 hCoV-19/USA/MD-MDH-0400/2020\|EPI_ISL_660094\|2020-1 | ···················· | ·A·················· | ···················· | ···················· | ···················· | ···················· |
| S | 2044 hCoV-19/USA/MD-MDH-0401/2020\|EPI_ISL_660095\|2020-1 | ···················· | ·A·················· | ···················· | ···················· | ···················· | ···················· |
| S | 2045 hCoV-19/USA/MD-MDH-0402/2020\|EPI_ISL_660096\|2020-1 | ···················· | ·A·················· | ···················· | ···················· | ···················· | ···················· |
| S | 2046 hCoV-19/USA/MD-MDH-0403/2020\|EPI_ISL_660097\|2020-1 | ···················· | ·A·················· | ···················· | ···················· | ···················· | ···················· |
| S | 2047 hCoV-19/USA/MD-MDH-0405/2020\|EPI_ISL_660098\|2020-1 | ···················· | ···················· | ···················· | ···················· | ···················· | ···················· |
| S | 2048 hCoV-19/USA/MD-MDH-0390/2020\|EPI_ISL_660099\|2020-1 | ···················· | ·A·················· | ···················· | ···················· | ···················· | ···················· |
| S | 2052 hCoV-19/England/QEUH-AAAC28/2020\|EPI_ISL_651103\|20 | ···················· | ·A·················· | ···················· | ···················· | ···················· | ··A················· |
| S | 2053 hCoV-19/England/QEUH-AAB44D/2020\|EPI_ISL_651104\|20 | ···················· | ·A·················· | ···················· | ···················· | ···················· | ··A················· |
| S | 2054 hCoV-19/England/QEUH-AAC78F/2020\|EPI_ISL_651105\|20 | ···················· | ·A·················· | ···················· | ···················· | ···················· | ··A················· |
| S | 2055 hCoV-19/England/QEUH-AAC822/2020\|EPI_ISL_651106\|20 | ···················· | ·A·················· | ···················· | ···················· | ···················· | ··A················· |
| S | 2056 hCoV-19/England/NOTT-11C6D1/2020\|EPI_ISL_651107\|20 | ···················· | ···················· | ···················· | ···················· | ···················· | ···················· |
| S | 2057 hCoV-19/Wales/PHWC-489D7D/2020\|EPI_ISL_651108\|2020 | ···················· | ···················· | ···················· | ···················· | ···················· | ···················· |
| S | 2058 hCoV-19/England/CAMC-B230EE/2020\|EPI_ISL_651109\|20 | ···················· | ·A·················· | ···················· | ···················· | ···················· | ··A················· |
| S | 2059 hCoV-19/England/NOTT-11C1B2/2020\|EPI_ISL_652400\|20 | ···················· | ···················· | ···················· | ···················· | ···················· | ···················· |
| S | 2060 hCoV-19/England/NOTT-11C2FB/2020\|EPI_ISL_652401\|20 | ···················· | ···················· | ···················· | ···················· | ···················· | ···················· |
| S | 2061 hCoV-19/England/NOTT-11C334/2020\|EPI_ISL_652402\|20 | ···················· | ···················· | ···················· | ···················· | ···················· | ···················· |
| S | 2062 hCoV-19/England/NOTT-11C361/2020\|EPI_ISL_652403\|20 | ···················· | ···················· | ···················· | ···················· | ···················· | ···················· |
| S | 2063 hCoV-19/England/NOTT-11C370/2020\|EPI_ISL_652404\|20 | ···················· | ···················· | ···················· | ···················· | ···················· | ···················· |
| S | 2064 hCoV-19/England/NOTT-11C6C2/2020\|EPI_ISL_652407\|20 | ···················· | ···················· | ···················· | ···················· | ···················· | ···················· |
| S | 2065 hCoV-19/England/NOTT-11C7B0/2020\|EPI_ISL_652410\|20 | ···················· | ···················· | ···················· | ···················· | ···················· | ···················· |
| S | 2066 hCoV-19/England/NOTT-11C7ED/2020\|EPI_ISL_652411\|20 | ···················· | ···················· | ···················· | ···················· | ···················· | ···················· |
| S | 2067 hCoV-19/England/NOTT-11C826/2020\|EPI_ISL_652412\|20 | ···················· | ···················· | ···················· | ···················· | ···················· | ···················· |
| S | 2068 hCoV-19/England/NOTT-11C844/2020\|EPI_ISL_652413\|20 | ···················· | ···················· | ···················· | ···················· | ···················· | ···················· |
| S | 2069 hCoV-19/Wales/PHWC-489566/2020\|EPI_ISL_652414\|2020 | ···················· | ···················· | ···················· | ···················· | ···················· | ···················· |
| S | 2070 hCoV-19/Wales/PHWC-48985E/2020\|EPI_ISL_652415\|2020 | ···················· | ···················· | ···················· | ···················· | ···················· | ···················· |
| S | 2071 hCoV-19/Wales/PHWC-48987C/2020\|EPI_ISL_652416\|2020 | ···················· | ···················· | ···················· | ···················· | ···················· | ···················· |
| S | 2072 hCoV-19/Wales/PHWC-48988B/2020\|EPI_ISL_652417\|2020 | ···················· | ···················· | ···················· | ···················· | ···················· | ···················· |
| S | 2073 hCoV-19/Wales/PHWC-48989A/2020\|EPI_ISL_652418\|2020 | ···················· | ···················· | ···················· | ···················· | ···················· | ···················· |
| S | 2074 hCoV-19/Wales/PHWC-4898A9/2020\|EPI_ISL_652419\|2020 | ···················· | ···················· | ···················· | ···················· | ···················· | ···················· |
| S | 2075 hCoV-19/Wales/PHWC-4898B8/2020\|EPI_ISL_652420\|2020 | ···················· | ···················· | ···················· | ···················· | ···················· | ···················· |
| S | 2076 hCoV-19/Wales/PHWC-4898C7/2020\|EPI_ISL_652421\|2020 | ···················· | ···················· | ···················· | ···················· | ···················· | ···················· |

| | | | | | | | |
|---|---|---|---|---|---|---|---|
| S | 2077 hCoV-19/Wales/PHWC-4898D6/2020\|EPI_ISL_652422\|2020 | ···················· | ···················· | ····················· | ···················· | ···················· | ···················· |
| S | 2078 hCoV-19/Wales/PHWC-4898E5/2020\|EPI_ISL_652423\|2020 | ···················· | ···················· | ····················· | ···················· | ···················· | ···················· |
| S | 2079 hCoV-19/Wales/PHWC-489900/2020\|EPI_ISL_652424\|2020 | ···················· | ···················· | ····················· | ···················· | ···················· | ···················· |
| S | 2080 hCoV-19/Wales/PHWC-48991F/2020\|EPI_ISL_652425\|2020 | ···················· | ···················· | ····················· | ···················· | ···················· | ···················· |
| S | 2081 hCoV-19/Wales/PHWC-48992E/2020\|EPI_ISL_652426\|2020 | ···················· | ···················· | ····················· | ···················· | ···················· | ···················· |
| S | 2082 hCoV-19/Wales/PHWC-48993D/2020\|EPI_ISL_652427\|2020 | ···················· | ···················· | ····················· | ···················· | ···················· | ···················· |
| S | 2083 hCoV-19/Wales/PHWC-48994C/2020\|EPI_ISL_652428\|2020 | ···················· | ···················· | ····················· | ···················· | ···················· | ···················· |
| S | 2084 hCoV-19/Wales/PHWC-48995B/2020\|EPI_ISL_652429\|2020 | ···················· | ···················· | ····················· | ···················· | ···················· | ···················· |
| S | 2085 hCoV-19/Wales/PHWC-48996A/2020\|EPI_ISL_652430\|2020 | ···················· | ···················· | ····················· | ···················· | ···················· | ···················· |
| S | 2086 hCoV-19/Wales/PHWC-489988/2020\|EPI_ISL_652431\|2020 | ···················· | ···················· | ····················· | ···················· | ···················· | ···················· |
| S | 2087 hCoV-19/Wales/PHWC-489997/2020\|EPI_ISL_652432\|2020 | ···················· | ···················· | ····················· | ···················· | ···················· | ···················· |
| S | 2088 hCoV-19/Wales/PHWC-4899A6/2020\|EPI_ISL_652433\|2020 | ···················· | ···················· | ····················· | ···················· | ···················· | ···················· |
| S | 2089 hCoV-19/Wales/PHWC-4899B5/2020\|EPI_ISL_652434\|2020 | ···················· | ···················· | ····················· | ···················· | ···················· | ···················· |
| S | 2090 hCoV-19/Wales/PHWC-4899C4/2020\|EPI_ISL_652435\|2020 | ···················· | ···················· | ····················· | ···················· | ···················· | ···················· |
| S | 2091 hCoV-19/Wales/PHWC-4899D3/2020\|EPI_ISL_652436\|2020 | ···················· | ···················· | ····················· | ···················· | ···················· | ···················· |
| S | 2092 hCoV-19/Wales/PHWC-4899E2/2020\|EPI_ISL_652437\|2020 | ···················· | ···················· | ····················· | ···················· | ···················· | ···················· |
| S | 2093 hCoV-19/Wales/PHWC-489A0D/2020\|EPI_ISL_652438\|2020 | ···················· | ···················· | ····················· | ···················· | ···················· | ···················· |
| S | 2094 hCoV-19/Wales/PHWC-489A1C/2020\|EPI_ISL_652439\|2020 | ···················· | ···················· | ····················· | ···················· | ···················· | ···················· |
| S | 2095 hCoV-19/Wales/PHWC-489A2B/2020\|EPI_ISL_652440\|2020 | ···················· | ···················· | ····················· | ···················· | ···················· | ···················· |
| S | 2096 hCoV-19/Wales/PHWC-489A3A/2020\|EPI_ISL_652441\|2020 | ···················· | ···················· | ····················· | ···················· | ···················· | ···················· |
| S | 2097 hCoV-19/Wales/PHWC-489A49/2020\|EPI_ISL_652442\|2020 | ···················· | ···················· | ····················· | ···················· | ···················· | ···················· |
| S | 2098 hCoV-19/Wales/PHWC-489A58/2020\|EPI_ISL_652443\|2020 | ···················· | ···················· | ····················· | ···················· | ···················· | ···················· |
| S | 2099 hCoV-19/Wales/PHWC-489A67/2020\|EPI_ISL_652444\|2020 | ···················· | ···················· | ····················· | ···················· | ···················· | ···················· |
| S | 2100 hCoV-19/Wales/PHWC-489A76/2020\|EPI_ISL_652445\|2020 | ···················· | ···················· | ····················· | ···················· | ···················· | ···················· |
| S | 2101 hCoV-19/Wales/PHWC-489A85/2020\|EPI_ISL_652446\|2020 | ···················· | ···················· | ····················· | ···················· | ···················· | ···················· |
| S | 2102 hCoV-19/Wales/PHWC-489A94/2020\|EPI_ISL_652447\|2020 | ···················· | ···················· | ····················· | ···················· | ···················· | ···················· |
| S | 2103 hCoV-19/Wales/PHWC-489AA3/2020\|EPI_ISL_652448\|2020 | ···················· | ···················· | ····················· | ···················· | ···················· | ···················· |
| S | 2104 hCoV-19/Wales/PHWC-489AB2/2020\|EPI_ISL_652449\|2020 | ···················· | ···················· | ····················· | ···················· | ···················· | ···················· |
| S | 2105 hCoV-19/Wales/PHWC-489AC1/2020\|EPI_ISL_652450\|2020 | ···················· | ···················· | ····················· | ···················· | ···················· | ···················· |
| S | 2106 hCoV-19/Wales/PHWC-489AD0/2020\|EPI_ISL_652451\|2020 | ···················· | ···················· | ····················· | ···················· | ···················· | ···················· |
| S | 2107 hCoV-19/Wales/PHWC-489AEF/2020\|EPI_ISL_652452\|2020 | ···················· | ···················· | ····················· | ···················· | ···················· | ···················· |
| S | 2108 hCoV-19/Wales/PHWC-489AFE/2020\|EPI_ISL_652453\|2020 | ···················· | ···················· | ····················· | ···················· | ···················· | ···················· |
| S | 2109 hCoV-19/Wales/PHWC-489B0A/2020\|EPI_ISL_652454\|2020 | ···················· | ···················· | ····················· | ···················· | ···················· | ···················· |
| S | 2110 hCoV-19/Wales/PHWC-489B19/2020\|EPI_ISL_652455\|2020 | ···················· | ···················· | ····················· | ···················· | ···················· | ···················· |
| S | 2111 hCoV-19/USA/LA-EVTL1030/2020\|EPI_ISL_653721\|2020-0 | ···················· | ···················· | ····················· | ···················· | ···················· | ···················· |
| S | 2112 hCoV-19/USA/LA-EVTL1032/2020\|EPI_ISL_653722\|2020-0 | ···················· | ···················· | ····················· | ···················· | ···················· | ···················· |
| S | 2113 hCoV-19/USA/LA-EVTL1033/2020\|EPI_ISL_653723\|2020-0 | ···················· | ···················· | ····················· | ···················· | ···················· | ···················· |
| S | 2114 hCoV-19/USA/LA-EVTL1034/2020\|EPI_ISL_653724\|2020-0 | ···················· | ···················· | ····················· | ···················· | ···················· | ···················· |
| S | 2115 hCoV-19/USA/LA-EVTL1035/2020\|EPI_ISL_653725\|2020-0 | ···················· | ···················· | ····················· | ···················· | ···················· | ···················· |
| S | 2116 hCoV-19/USA/LA-EVTL1036/2020\|EPI_ISL_653726\|2020-0 | ···················· | ···················· | ····················· | ···················· | ···················· | ···················· |
| S | 2117 hCoV-19/USA/LA-EVTL1037/2020\|EPI_ISL_653727\|2020-0 | ···················· | ···················· | ····················· | ···················· | ···················· | ···················· |
| S | 2118 hCoV-19/USA/LA-EVTL1038/2020\|EPI_ISL_653728\|2020-0 | ···················· | ···················· | ····················· | ···················· | ···················· | ···················· |
| S | 2119 hCoV-19/USA/LA-EVTL1039/2020\|EPI_ISL_653729\|2020-0 | ···················· | ···················· | ····················· | ···················· | ···················· | ···················· |
| S | 2120 hCoV-19/USA/LA-EVTL1040/2020\|EPI_ISL_653730\|2020-0 | ···················· | ···················· | ····················· | ···················· | ···················· | ···················· |
| S | 2121 hCoV-19/USA/LA-EVTL1041/2020\|EPI_ISL_653731\|2020-0 | ···················· | ···················· | ····················· | ···················· | ···················· | ···················· |
| S | 2122 hCoV-19/USA/LA-EVTL1042/2020\|EPI_ISL_653732\|2020-0 | ···················· | ···················· | ····················· | ···················· | ···················· | ···················· |
| S | 2123 hCoV-19/USA/LA-EVTL1043/2020\|EPI_ISL_653733\|2020-0 | ···················· | ···················· | ····················· | ···················· | ···················· | ···················· |
| S | 2124 hCoV-19/USA/LA-EVTL1044/2020\|EPI_ISL_653734\|2020-0 | ···················· | ···················· | ····················· | ···················· | ···················· | ···················· |
| S | 2125 hCoV-19/USA/LA-EVTL1045/2020\|EPI_ISL_653735\|2020-0 | ···················· | ···················· | ····················· | ···················· | ···················· | ···················· |
| S | 2126 hCoV-19/USA/LA-EVTL1048/2020\|EPI_ISL_653738\|2020-0 | ···················· | ···················· | ····················· | ···················· | ···················· | ···················· |
| S | 2127 hCoV-19/USA/LA-EVTL1049/2020\|EPI_ISL_653739\|2020-0 | ···················· | ···················· | ····················· | ···················· | ···················· | ···················· |
| S | 2128 hCoV-19/USA/LA-EVTL1050/2020\|EPI_ISL_653740\|2020-0 | ···················· | ···················· | ····················· | ···················· | ···················· | ···················· |
| S | 2129 hCoV-19/USA/LA-EVTL1051/2020\|EPI_ISL_653741\|2020-0 | ···················· | ·············A······ | ····················· | ···················· | ···················· | ···················· |
| S | 2130 hCoV-19/USA/LA-EVTL1052/2020\|EPI_ISL_653742\|2020-0 | ···················· | ···················· | ····················· | ···················· | ···················· | ···················· |
| S | 2131 hCoV-19/USA/LA-EVTL1053/2020\|EPI_ISL_653743\|2020-0 | ···················· | ···················· | ····················· | ···················· | ···················· | ···················· |
| S | 2132 hCoV-19/USA/LA-EVTL1054/2020\|EPI_ISL_653744\|2020-0 | ···················· | ···················· | ····················· | ···················· | ···················· | ···················· |
| S | 2135 hCoV-19/Colombia/DC-INS-84587/2020\|EPI_ISL_653750\| | ···················· | ···················· | ····················· | ···················· | ···················· | ···················· |
| S | 2136 hCoV-19/Colombia/VAC-INS-82835/2020\|EPI_ISL_653751 | ···················· | ···················· | ····················· | ···················· | ···················· | ···················· |
| S | 2137 hCoV-19/Colombia/NSA-NS-79852/2020\|EPI_ISL_653752\| | ···················· | ···················· | ····················· | ···················· | ···················· | ···················· |
| S | 2138 hCoV-19/Colombia/DC-INS-3334/2020\|EPI_ISL_653754\|2 | ···················· | ···················· | ····················· | ···················· | ···················· | ···················· |
| S | 2140 hCoV-19/Colombia/ANT-INS-01/2020\|EPI_ISL_653756\|20 | ···················· | ···················· | ····················· | ···················· | ···················· | ···················· |
| S | 2142 hCoV-19/Italy/APU-IZSPB_198PT/2020\|EPI_ISL_653763\| | ···················· | ···················· | ····················· | ···················· | ···················· | ···················· |
| S | 2143 hCoV-19/Italy/APU-IZSPB_200PT/2020\|EPI_ISL_653764\| | ···················· | ···················· | ····················· | ······T············· | ···················· | ···················· |
| S | 2144 hCoV-19/Italy/APU-IZSPB_269PT/2020\|EPI_ISL_653765\| | ···················· | ···················· | ····················· | ···················· | ···················· | ···················· |
| S | 2145 hCoV-19/Italy/APU-IZSPB_271PT/2020\|EPI_ISL_653766\| | ···················· | ···················· | ····················· | ···················· | ···················· | ···················· |
| S | 2146 hCoV-19/Italy/APU-IZSPB_272PT/2020\|EPI_ISL_653767\| | ···················· | ···················· | ····················· | ···················· | ···················· | ···················· |
| S | 2147 hCoV-19/Italy/APU-IZSPB_274PT/2020\|EPI_ISL_653768\| | ···················· | ···················· | ····················· | ···················· | ···················· | ···················· |
| S | 2148 hCoV-19/Italy/APU-IZSPB_275PT/2020\|EPI_ISL_653769\| | ···················· | ···················· | ····················· | ···················· | ······T············· | ···················· |
| S | 2149 hCoV-19/Italy/APU-IZSPB_276PT/2020\|EPI_ISL_653770\| | ···················· | ···················· | ····················· | ···················· | ···················· | ···················· |
| S | 2150 hCoV-19/Italy/APU-IZSPB_277PT/2020\|EPI_ISL_653771\| | ···················· | ···················· | ····················· | ···················· | ···················· | ···················· |
| S | 2151 hCoV-19/Italy/APU-IZSPB_278PT/2020\|EPI_ISL_653772\| | ···················· | ···················· | ····················· | ···················· | ···················· | ···················· |
| S | 2152 hCoV-19/Italy/APU-IZSPB_280PT/2020\|EPI_ISL_653773\| | ···················· | ···················· | ····················· | ···················· | ······T············· | ···················· |
| S | 2153 hCoV-19/Italy/APU-IZSPB_281PT/2020\|EPI_ISL_653774\| | ···················· | ···················· | ····················· | ···················· | ······T············· | ···················· |
| S | 2154 hCoV-19/Italy/APU-IZSPB_282PT/2020\|EPI_ISL_653775\| | ···················· | ···················· | ····················· | ···················· | ···················· | ···················· |
| S | 2155 hCoV-19/Italy/APU-IZSPB_284PT/2020\|EPI_ISL_653776\| | ···················· | ···················· | ····················· | ···················· | ···················· | ···················· |
| S | 2156 hCoV-19/Italy/APU-IZSPB_285PT/2020\|EPI_ISL_653777\| | ···················· | ···················· | ····················· | ···················· | ···················· | ···················· |
| S | 2157 hCoV-19/Italy/APU-IZSPB_286PT/2020\|EPI_ISL_653778\| | ···················· | ···················· | ····················· | ···················· | ···················· | ···················· |
| S | 2158 hCoV-19/Italy/APU-IZSPB_287PT/2020\|EPI_ISL_653779\| | ···················· | ···················· | ····················· | ···················· | ······T············· | ···················· |
| S | 2159 hCoV-19/Italy/APU-IZSPB_289PT/2020\|EPI_ISL_653780\| | ···················· | ·A·················· | ····················· | ···················· | ···················· | ···················· |
| S | 2160 hCoV-19/Italy/APU-IZSPB_290PT/2020\|EPI_ISL_653781\| | ···················· | ···················· | ····················· | ···················· | ···················· | ···················· |

| | | | | | | | | |
|---|---|---|---|---|---|---|---|---|
| S | 2161 | hCoV-19/Italy/APU-IZSPB_291PT/2020\|EPI_ISL_653782\| | ···················· | ···················· | ····················· | ···················· | ···················· | ···················· |
| S | 2162 | hCoV-19/Italy/APU-IZSPB_292PT/2020\|EPI_ISL_653783\| | ···················· | ···················· | ····················· | ···················· | ···················· | ···················· |
| S | 2163 | hCoV-19/Italy/APU-IZSPB_265PT/2020\|EPI_ISL_653784\| | ···················· | ···················· | ····················· | ···················· | ···················· | ···················· |
| S | 2164 | hCoV-19/Italy/APU-IZSPB_266PT/2020\|EPI_ISL_653785\| | ···················· | ···················· | ····················· | ···················· | ···················· | ···················· |
| S | 2165 | hCoV-19/Italy/APU-IZSPB_267PT/2020\|EPI_ISL_653786\| | ···················· | ···················· | ····················· | ···················· | ···················· | ···················· |
| S | 2166 | hCoV-19/England/ALDP-B5B056/2020\|EPI_ISL_655587\|20 | ···················· | ···················· | ····················· | ···················· | ···················· | ···················· |
| S | 2167 | hCoV-19/England/ALDP-B63D27/2020\|EPI_ISL_655588\|20 | ···················· | ···················· | ····················· | ···················· | ···················· | ···················· |
| S | 2168 | hCoV-19/England/QEUH-B42465/2020\|EPI_ISL_657230\|20 | ···················· | ···················· | ····················· | ···················· | ···················· | ···················· |
| S | 2169 | hCoV-19/England/QEUH-B42492/2020\|EPI_ISL_657231\|20 | ···················· | ···················· | ····················· | ···················· | ···················· | ···················· |
| S | 2170 | hCoV-19/England/QEUH-B43516/2020\|EPI_ISL_657232\|20 | ···················· | ···················· | ····················· | ···················· | ···················· | ···················· |
| S | 2171 | hCoV-19/England/QEUH-B404D0/2020\|EPI_ISL_657233\|20 | ···················· | ···················· | ····················· | ···················· | ···················· | ···················· |
| S | 2172 | hCoV-19/England/QEUH-B421C8/2020\|EPI_ISL_657234\|20 | ···················· | ···················· | ····················· | ···················· | ·····T·············· | ···················· |
| S | 2173 | hCoV-19/England/QEUH-B3FFB5/2020\|EPI_ISL_657235\|20 | ···················· | ···················· | ····················· | ···················· | ···················· | ···················· |
| S | 2174 | hCoV-19/England/QEUH-B425EA/2020\|EPI_ISL_657237\|20 | ···················· | ···················· | ····················· | ···················· | ···················· | ···················· |
| S | 2175 | hCoV-19/England/QEUH-B438E0/2020\|EPI_ISL_657238\|20 | ···················· | ···················· | ····················· | ···················· | ···················· | ···················· |
| S | 2176 | hCoV-19/England/QEUH-B435F8/2020\|EPI_ISL_657239\|20 | ···················· | ···················· | ····················· | ···················· | ···················· | ···················· |
| S | 2177 | hCoV-19/England/QEUH-B4217D/2020\|EPI_ISL_657240\|20 | ···················· | ···················· | ····················· | ···················· | ···················· | ···················· |
| S | 2178 | hCoV-19/England/QEUH-B40230/2020\|EPI_ISL_657242\|20 | ···················· | ···················· | ····················· | ···················· | ···················· | ···················· |
| S | 2179 | hCoV-19/England/QEUH-B40537/2020\|EPI_ISL_657243\|20 | ···················· | ···················· | ····················· | ···················· | ···················· | ···················· |
| S | 2180 | hCoV-19/England/QEUH-B4069E/2020\|EPI_ISL_657244\|20 | ···················· | ···················· | ····················· | ···················· | ···················· | ···················· |
| S | 2181 | hCoV-19/England/QEUH-B43543/2020\|EPI_ISL_657245\|20 | ···················· | ···················· | ····················· | ···················· | ···················· | ···················· |
| S | 2182 | hCoV-19/England/QEUH-B4076E/2020\|EPI_ISL_657246\|20 | ···················· | ···················· | ····················· | ···················· | ···················· | ···················· |
| S | 2183 | hCoV-19/England/QEUH-B4273F/2020\|EPI_ISL_657247\|20 | ···················· | ·A·················· | ····················· | ···················· | ···················· | ··A················· |
| S | 2184 | hCoV-19/England/QEUH-B3FFA6/2020\|EPI_ISL_657248\|20 | ···················· | ···················· | ····················· | ···················· | ···················· | ···················· |
| S | 2185 | hCoV-19/England/QEUH-B409B3/2020\|EPI_ISL_657249\|20 | ···················· | ···················· | ····················· | ···················· | ···················· | ···················· |
| S | 2186 | hCoV-19/England/QEUH-B42210/2020\|EPI_ISL_657250\|20 | ···················· | ···················· | ····················· | ···················· | ···················· | ···················· |
| S | 2187 | hCoV-19/England/QEUH-B4332B/2020\|EPI_ISL_657251\|20 | ···················· | ···················· | ····················· | ···················· | ···················· | ···················· |
| S | 2188 | hCoV-19/England/QEUH-B43789/2020\|EPI_ISL_657252\|20 | ···················· | ···················· | ····················· | ···················· | ···················· | ···················· |
| S | 2189 | hCoV-19/England/QEUH-B43622/2020\|EPI_ISL_657253\|20 | ···················· | ···················· | ····················· | ···················· | ···················· | ···················· |
| S | 2190 | hCoV-19/England/QEUH-B4077D/2020\|EPI_ISL_657254\|20 | ···················· | ···················· | ····················· | ···················· | ···················· | ···················· |
| S | 2191 | hCoV-19/England/QEUH-B4377A/2020\|EPI_ISL_657256\|20 | ···················· | ···················· | ····················· | ···················· | ···················· | ···················· |
| S | 2192 | hCoV-19/England/QEUH-B4226B/2020\|EPI_ISL_657258\|20 | ···················· | ···················· | ····················· | ···················· | ···················· | ···················· |
| S | 2193 | hCoV-19/England/QEUH-B40485/2020\|EPI_ISL_657259\|20 | ···················· | ·A·················· | ····················· | ···················· | ···················· | ··A················· |
| S | 2194 | hCoV-19/England/QEUH-B406E9/2020\|EPI_ISL_657260\|20 | ···················· | ···················· | ····················· | ···················· | ···················· | ···················· |
| S | 2195 | hCoV-19/England/QEUH-B4340A/2020\|EPI_ISL_657264\|20 | ···················· | ···················· | ····················· | ···················· | ···················· | ···················· |
| S | 2196 | hCoV-19/England/QEUH-B4277B/2020\|EPI_ISL_657265\|20 | ···················· | ···················· | ····················· | ···················· | ···················· | ···················· |
| S | 2197 | hCoV-19/England/QEUH-B40661/2020\|EPI_ISL_657266\|20 | ···················· | ···················· | ····················· | ···················· | ···················· | ···················· |
| S | 2198 | hCoV-19/England/QEUH-B422F2/2020\|EPI_ISL_657267\|20 | ···················· | ···················· | ····················· | ···················· | ···················· | ···················· |
| S | 2199 | hCoV-19/England/QEUH-B4017F/2020\|EPI_ISL_657268\|20 | ···················· | ···················· | ····················· | ···················· | ···················· | ···················· |
| S | 2200 | hCoV-19/England/QEUH-B4085C/2020\|EPI_ISL_657269\|20 | ···················· | ···················· | ····················· | ···················· | ···················· | ···················· |
| S | 2201 | hCoV-19/England/QEUH-B40959/2020\|EPI_ISL_657270\|20 | ···················· | ···················· | ····················· | ···················· | ···················· | ···················· |
| S | 2202 | hCoV-19/England/QEUH-B42641/2020\|EPI_ISL_657271\|20 | ···················· | ···················· | ····················· | ···················· | ···················· | ···················· |
| S | 2203 | hCoV-19/England/QEUH-B4093B/2020\|EPI_ISL_657272\|20 | ···················· | ···················· | ····················· | ···················· | ···················· | ···················· |
| S | 2204 | hCoV-19/England/QEUH-B40115/2020\|EPI_ISL_657273\|20 | ···················· | ···················· | ····················· | ···················· | ···················· | ···················· |
| S | 2205 | hCoV-19/England/QEUH-B424FC/2020\|EPI_ISL_657274\|20 | ···················· | ···················· | ····················· | ···················· | ···················· | ···················· |
| S | 2206 | hCoV-19/England/QEUH-B4018E/2020\|EPI_ISL_657275\|20 | ···················· | ·A·················· | ····················· | ···················· | ···················· | ···················· |
| S | 2207 | hCoV-19/England/QEUH-B4372F/2020\|EPI_ISL_657276\|20 | ···················· | ···················· | ····················· | ···················· | ···················· | ···················· |
| S | 2208 | hCoV-19/England/QEUH-B405BF/2020\|EPI_ISL_657277\|20 | ···················· | ···················· | ····················· | ···················· | ···················· | ···················· |
| S | 2209 | hCoV-19/England/QEUH-B401AC/2020\|EPI_ISL_657278\|20 | ···················· | ···················· | ····················· | ···················· | ···················· | ···················· |
| S | 2210 | hCoV-19/England/QEUH-B43859/2020\|EPI_ISL_657279\|20 | ···················· | ···················· | ····················· | ···················· | ···················· | ···················· |
| S | 2211 | hCoV-19/England/QEUH-B4043A/2020\|EPI_ISL_657280\|20 | ···················· | ···················· | ····················· | ···················· | ···················· | ···················· |
| S | 2212 | hCoV-19/England/QEUH-B4028B/2020\|EPI_ISL_657281\|20 | ···················· | ·A·················· | ····················· | ···················· | ···················· | ··A················· |
| S | 2213 | hCoV-19/England/QEUH-B4333A/2020\|EPI_ISL_657282\|20 | ···················· | ···················· | ····················· | ···················· | ···················· | ···················· |
| S | 2214 | hCoV-19/England/QEUH-B435CB/2020\|EPI_ISL_657283\|20 | ···················· | ···················· | ····················· | ···················· | ···················· | ···················· |
| S | 2215 | hCoV-19/England/QEUH-B434BF/2020\|EPI_ISL_657284\|20 | ···················· | ···················· | ····················· | ···················· | ···················· | ···················· |
| S | 2216 | hCoV-19/England/QEUH-B43446/2020\|EPI_ISL_657285\|20 | ···················· | ···················· | ····················· | ···················· | ···················· | ···················· |
| S | 2217 | hCoV-19/England/QEUH-B3FF88/2020\|EPI_ISL_657286\|20 | ···················· | ···················· | ····················· | ···················· | ···················· | ···················· |
| S | 2218 | hCoV-19/England/QEUH-B3FF79/2020\|EPI_ISL_657287\|20 | ···················· | ···················· | ····················· | ···················· | ···················· | ···················· |
| S | 2219 | hCoV-19/England/QEUH-B436F5/2020\|EPI_ISL_657288\|20 | ···················· | ···················· | ····················· | ···················· | ···················· | ···················· |
| S | 2220 | hCoV-19/England/QEUH-B3FE8B/2020\|EPI_ISL_657289\|20 | ···················· | ···················· | ····················· | ···················· | ···················· | ···················· |
| S | 2221 | hCoV-19/England/QEUH-B4358F/2020\|EPI_ISL_657290\|20 | ···················· | ···················· | ····················· | ···················· | ···················· | ···················· |
| S | 2222 | hCoV-19/England/QEUH-B42131/2020\|EPI_ISL_657291\|20 | ···················· | ···················· | ····················· | ···················· | ···················· | ···················· |
| S | 2223 | hCoV-19/England/QEUH-B4029A/2020\|EPI_ISL_657292\|20 | ···················· | ···················· | ····················· | ···················· | ···················· | ···················· |
| S | 2224 | hCoV-19/England/QEUH-B435DA/2020\|EPI_ISL_657293\|20 | ···················· | ···················· | ····················· | ···················· | ···················· | ···················· |
| S | 2225 | hCoV-19/England/QEUH-B40221/2020\|EPI_ISL_657296\|20 | ···················· | ···················· | ····················· | ···················· | ···················· | ···················· |
| S | 2226 | hCoV-19/England/QEUH-B40616/2020\|EPI_ISL_657297\|20 | ···················· | ···················· | ····················· | ···················· | ···················· | ···················· |
| S | 2227 | hCoV-19/England/QEUH-B434CE/2020\|EPI_ISL_657298\|20 | ···················· | ···················· | ····················· | ···················· | ···················· | ···················· |
| S | 2228 | hCoV-19/England/QEUH-B422E3/2020\|EPI_ISL_657299\|20 | ···················· | ···················· | ····················· | ···················· | ···················· | ···················· |
| S | 2229 | hCoV-19/England/QEUH-B3FCCD/2020\|EPI_ISL_658560\|20 | ···················· | ···················· | ····················· | ···················· | ···················· | ···················· |
| S | 2230 | hCoV-19/England/QEUH-B3F9C6/2020\|EPI_ISL_658561\|20 | ···················· | ···················· | ····················· | ···················· | ···················· | ···················· |
| S | 2231 | hCoV-19/England/QEUH-B414C0/2020\|EPI_ISL_658562\|20 | ···················· | ···················· | ····················· | ···················· | ···················· | ···················· |
| S | 2232 | hCoV-19/England/QEUH-B3FAF0/2020\|EPI_ISL_658563\|20 | ···················· | ···················· | ····················· | ···················· | ···················· | ···················· |
| S | 2233 | hCoV-19/England/QEUH-B3FD9D/2020\|EPI_ISL_658564\|20 | ···················· | ···················· | ····················· | ···················· | ···················· | ···················· |
| S | 2234 | hCoV-19/England/QEUH-B41053/2020\|EPI_ISL_658565\|20 | ···················· | ···················· | ····················· | ···················· | ···················· | ···················· |
| S | 2235 | hCoV-19/England/QEUH-B3FD15/2020\|EPI_ISL_658566\|20 | ···················· | ···················· | ····················· | ···················· | ···················· | ···················· |
| S | 2236 | hCoV-19/England/QEUH-B3FAA5/2020\|EPI_ISL_658567\|20 | ···················· | ···················· | ····················· | ···················· | ···················· | ···················· |
| S | 2237 | hCoV-19/England/QEUH-B418C4/2020\|EPI_ISL_658568\|20 | ···················· | ···················· | ····················· | ···················· | ···················· | ···················· |
| S | 2238 | hCoV-19/England/QEUH-B415DC/2020\|EPI_ISL_658569\|20 | ···················· | ···················· | ····················· | ···················· | ···················· | ···················· |
| S | 2239 | hCoV-19/England/QEUH-B3F47A/2020\|EPI_ISL_658570\|20 | ···················· | ···················· | ····················· | ···················· | ···················· | ···················· |
| S | 2240 | hCoV-19/England/QEUH-B4193A/2020\|EPI_ISL_658571\|20 | ···················· | ···················· | ····················· | ···················· | ···················· | ···················· |

| | | | | | | | |
|---|---|---|---|---|---|---|---|
| S | 2241 hCoV-19/England/QEUH-B3FAC3/2020\|EPI_ISL_658572\|20 | ···················· | ···················· | ····················· | ···················· | ···················· | ···················· |
| S | 2242 hCoV-19/England/QEUH-B3FB93/2020\|EPI_ISL_658573\|20 | ···················· | ···················· | ····················· | ···················· | ···················· | ···················· |
| S | 2243 hCoV-19/England/QEUH-B3F2BC/2020\|EPI_ISL_658574\|20 | ···················· | ···················· | ····················· | ···················· | ···················· | ···················· |
| S | 2244 hCoV-19/England/QEUH-B4141B/2020\|EPI_ISL_658575\|20 | ···················· | ···················· | ····················· | ···················· | ···················· | ···················· |
| S | 2245 hCoV-19/England/QEUH-B411C9/2020\|EPI_ISL_658577\|20 | ···················· | ···················· | ····················· | ···················· | ···················· | ···················· |
| S | 2246 hCoV-19/England/QEUH-B3F252/2020\|EPI_ISL_658578\|20 | ···················· | ···················· | ····················· | ···················· | ···················· | ···················· |
| S | 2247 hCoV-19/England/QEUH-B413E1/2020\|EPI_ISL_658579\|20 | ···················· | ···················· | ····················· | ···················· | ···················· | ···················· |
| S | 2248 hCoV-19/England/QEUH-B3F6FC/2020\|EPI_ISL_658580\|20 | ···················· | ·A·················· | ····················· | ···················· | ···················· | ···················· |
| S | 2249 hCoV-19/England/QEUH-B41396/2020\|EPI_ISL_658581\|20 | ···················· | ···················· | ····················· | ···················· | ···················· | ···················· |
| S | 2250 hCoV-19/England/QEUH-B41AEC/2020\|EPI_ISL_658582\|20 | ···················· | ···················· | ····················· | ···················· | ···················· | ···················· |
| S | 2251 hCoV-19/England/QEUH-B3F6DE/2020\|EPI_ISL_658583\|20 | ···················· | ·A·················· | ····················· | ···················· | ···················· | ···················· |
| S | 2252 hCoV-19/England/QEUH-B3F762/2020\|EPI_ISL_658585\|20 | ···················· | ·A·················· | ····················· | ···················· | ···················· | ···················· |
| S | 2253 hCoV-19/England/QEUH-B3F735/2020\|EPI_ISL_658586\|20 | ···················· | ···················· | ····················· | ···················· | ···················· | ···················· |
| S | 2254 hCoV-19/England/QEUH-B4109F/2020\|EPI_ISL_658587\|20 | ···················· | ···················· | ····················· | ···················· | ···················· | ···················· |
| S | 2255 hCoV-19/England/QEUH-B3F340/2020\|EPI_ISL_658588\|20 | ···················· | ···················· | ····················· | ···················· | ···················· | ···················· |
| S | 2256 hCoV-19/England/QEUH-B41044/2020\|EPI_ISL_658589\|20 | ···················· | ···················· | ····················· | ···················· | ···················· | ···················· |
| S | 2257 hCoV-19/England/QEUH-B3F234/2020\|EPI_ISL_658590\|20 | ···················· | ···················· | ····················· | ···················· | ···················· | ···················· |
| S | 2258 hCoV-19/England/QEUH-B3FBB1/2020\|EPI_ISL_658591\|20 | ···················· | ···················· | ····················· | ···················· | ···················· | ···················· |
| S | 2259 hCoV-19/England/QEUH-B413F0/2020\|EPI_ISL_658592\|20 | ···················· | ···················· | ····················· | ···················· | ···················· | ···················· |
| S | 2260 hCoV-19/England/QEUH-B41132/2020\|EPI_ISL_658593\|20 | ···················· | ···················· | ····················· | ···················· | ···················· | ···················· |
| S | 2261 hCoV-19/England/QEUH-B416BB/2020\|EPI_ISL_658594\|20 | ···················· | ···················· | ····················· | ···················· | ···················· | ···················· |
| S | 2262 hCoV-19/England/QEUH-B40FB1/2020\|EPI_ISL_658595\|20 | ···················· | ·A·················· | ····················· | ···················· | ···················· | ··A················· |
| S | 2263 hCoV-19/England/QEUH-B417E5/2020\|EPI_ISL_658596\|20 | ···················· | ···················· | ····················· | ···················· | ···················· | ···················· |
| S | 2264 hCoV-19/England/QEUH-B41879/2020\|EPI_ISL_658597\|20 | ···················· | ···················· | ····················· | ···················· | ···················· | ···················· |
| S | 2265 hCoV-19/England/QEUH-B414FD/2020\|EPI_ISL_658598\|20 | ···················· | ···················· | ····················· | ···················· | ···················· | ···················· |
| S | 2266 hCoV-19/England/QEUH-B303F4/2020\|EPI_ISL_659890\|20 | ···················· | ···················· | ····················· | ···················· | ···················· | ···················· |
| S | 2267 hCoV-19/England/QEUH-B2D5A9/2020\|EPI_ISL_659891\|20 | ···········T········ | ···················· | ····················· | ···················· | ···················· | ···················· |
| S | 2268 hCoV-19/England/QEUH-B2D65B/2020\|EPI_ISL_659892\|20 | ···················· | ···················· | ····················· | ···················· | ···················· | ···················· |
| S | 2269 hCoV-19/England/QEUH-B2F923/2020\|EPI_ISL_659893\|20 | ···················· | ···················· | ····················· | ···················· | ···················· | ···················· |
| S | 2270 hCoV-19/England/QEUH-B2EEDA/2020\|EPI_ISL_659894\|20 | ···················· | ···················· | ····················· | ···················· | ···················· | ···················· |
| S | 2271 hCoV-19/England/QEUH-B30B56/2020\|EPI_ISL_659895\|20 | ···················· | ···················· | ····················· | ···················· | ···················· | ···················· |
| S | 2272 hCoV-19/England/QEUH-B2D503/2020\|EPI_ISL_659896\|20 | ···················· | ···················· | ····················· | ···················· | ···················· | ···················· |
| S | 2273 hCoV-19/England/QEUH-B2D54F/2020\|EPI_ISL_659897\|20 | ···················· | ·A·················· | ····················· | ···················· | ···················· | ···················· |
| S | 2274 hCoV-19/England/QEUH-B30B74/2020\|EPI_ISL_659898\|20 | ···················· | ···················· | ····················· | ···················· | ···················· | ···················· |
| S | 2275 hCoV-19/England/QEUH-B2EAD6/2020\|EPI_ISL_659899\|20 | ···················· | ···················· | ····················· | ···················· | ···················· | ···················· |
| S | 2276 hCoV-19/USA/WI-GMF-23497/2020\|EPI_ISL_660871\|2020- | ···················· | ···················· | ····················· | ···················· | ···················· | ···················· |
| S | 2277 hCoV-19/USA/WI-GMF-23535/2020\|EPI_ISL_660872\|2020- | ···················· | ···················· | ····················· | ···················· | ···················· | ···················· |
| S | 2278 hCoV-19/USA/WI-GMF-23545/2020\|EPI_ISL_660873\|2020- | ···················· | ···················· | ····················· | ···················· | ···················· | ···················· |
| S | 2279 hCoV-19/USA/WI-GMF-23546/2020\|EPI_ISL_660874\|2020- | ···················· | ···················· | ····················· | ···················· | ···················· | ···················· |
| S | 2280 hCoV-19/USA/WI-GMF-23569/2020\|EPI_ISL_660875\|2020- | ···················· | ·A·················· | ····················· | ···················· | ···················· | ···················· |
| S | 2281 hCoV-19/USA/IA-GMF-23570/2020\|EPI_ISL_660876\|2020- | ···················· | ···················· | ····················· | ···················· | ···················· | ···················· |
| S | 2282 hCoV-19/USA/WI-GMF-23602/2020\|EPI_ISL_660877\|2020- | ···················· | ···················· | ····················· | ···················· | ···················· | ···················· |
| S | 2283 hCoV-19/USA/WI-GMF-23618/2020\|EPI_ISL_660878\|2020- | ···················· | ···················· | ····················· | ···················· | ···················· | ···················· |
| S | 2284 hCoV-19/USA/IA-GMF-23647/2020\|EPI_ISL_660879\|2020- | ···················· | ···················· | ····················· | ···················· | ···················· | ···················· |
| S | 2285 hCoV-19/USA/WI-GMF-23662/2020\|EPI_ISL_660880\|2020- | ···················· | ···················· | ····················· | ···················· | ···················· | ···················· |
| S | 2286 hCoV-19/USA/WI-GMF-23744/2020\|EPI_ISL_660881\|2020- | ···················· | ···················· | ····················· | ···················· | ···················· | ···················· |
| S | 2287 hCoV-19/USA/WI-GMF-23835/2020\|EPI_ISL_660882\|2020- | ···················· | ···················· | ····················· | ···················· | ···················· | ···················· |
| S | 2288 hCoV-19/USA/WI-GMF-23942/2020\|EPI_ISL_660883\|2020- | ···················· | ···················· | ····················· | ···················· | ···················· | ···················· |
| S | 2289 hCoV-19/USA/WI-GMF-23997/2020\|EPI_ISL_660884\|2020- | ···················· | ···················· | ····················· | ···················· | ···················· | ···················· |
| S | 2290 hCoV-19/USA/WI-GMF-24003/2020\|EPI_ISL_660885\|2020- | ···················· | ···················· | ····················· | ···················· | ···················· | ···················· |
| S | 2291 hCoV-19/USA/WI-GMF-24022/2020\|EPI_ISL_660886\|2020- | ···················· | ···················· | ····················· | ···················· | ···················· | ···················· |
| S | 2292 hCoV-19/USA/WI-GMF-24025/2020\|EPI_ISL_660887\|2020- | ···················· | ···················· | ····················· | ···················· | ···················· | ···················· |
| S | 2293 hCoV-19/USA/WI-GMF-24039/2020\|EPI_ISL_660888\|2020- | ···················· | ···················· | ····················· | ···················· | ···················· | ···················· |
| S | 2294 hCoV-19/USA/WI-GMF-24056/2020\|EPI_ISL_660889\|2020- | ···················· | ···················· | ····················· | ···················· | ···················· | ···················· |
| S | 2295 hCoV-19/USA/WI-GMF-24082/2020\|EPI_ISL_660890\|2020- | ···················· | ···················· | ····················· | ···················· | ···················· | ···················· |
| S | 2296 hCoV-19/USA/WI-GMF-24108/2020\|EPI_ISL_660891\|2020- | ···················· | ···················· | ····················· | ···················· | ···················· | ···················· |
| S | 2297 hCoV-19/USA/WI-GMF-24124/2020\|EPI_ISL_660892\|2020- | ···················· | ···················· | ····················· | ···················· | ···················· | ···················· |
| S | 2298 hCoV-19/USA/WI-GMF-24185/2020\|EPI_ISL_660893\|2020- | ···················· | ···················· | ····················· | ···················· | ···················· | ···················· |
| S | 2299 hCoV-19/USA/WI-GMF-24197/2020\|EPI_ISL_660894\|2020- | ···················· | ···················· | ····················· | ···················· | ···················· | ···················· |
| S | 2300 hCoV-19/USA/WI-GMF-24201/2020\|EPI_ISL_660895\|2020- | ···················· | ···················· | ····················· | ···················· | ···················· | ···················· |
| S | 2301 hCoV-19/USA/WI-GMF-24205/2020\|EPI_ISL_660896\|2020- | ···················· | ···················· | ····················· | ···················· | ···················· | ···················· |
| S | 2302 hCoV-19/USA/WI-GMF-24209/2020\|EPI_ISL_660897\|2020- | ···················· | ···················· | ····················· | ···················· | ···················· | ···················· |
| S | 2303 hCoV-19/USA/WI-GMF-24211/2020\|EPI_ISL_660898\|2020- | ···················· | ···················· | ····················· | ···················· | ···················· | ···················· |
| S | 2304 hCoV-19/USA/WI-GMF-24217/2020\|EPI_ISL_660899\|2020- | ···················· | ···················· | ····················· | ···················· | ···················· | ···················· |
| S | 2305 hCoV-19/England/QEUH-AABD42/2020\|EPI_ISL_651086\|20 | ···················· | ···················· | ····················· | ···················· | ···················· | ···················· |
| S | 2306 hCoV-19/England/QEUH-AAB36E/2020\|EPI_ISL_651087\|20 | ···················· | ·A·················· | ····················· | ···················· | ···················· | ··A················· |
| S | 2307 hCoV-19/England/QEUH-AAB51D/2020\|EPI_ISL_651088\|20 | ···················· | ···················· | ····················· | ···················· | ···················· | ···················· |
| S | 2308 hCoV-19/England/MILK-B05BD2/2020\|EPI_ISL_651089\|20 | ···················· | ···················· | ····················· | ···················· | ···················· | ···················· |
| S | 2309 hCoV-19/Northern Ireland/NIRE-10959E/2020\|EPI_ISL_ | ···················· | ···················· | ····················· | ···················· | ···················· | ···················· |
| S | 2310 hCoV-19/Northern Ireland/NIRE-1095CB/2020\|EPI_ISL_ | ···················· | ···················· | ····················· | ···················· | ···················· | ···················· |
| S | 2311 hCoV-19/England/NORT-286D45/2020\|EPI_ISL_652337\|20 | ···················· | ···················· | ····················· | ···················· | ···················· | ···················· |
| S | 2312 hCoV-19/England/NORT-286D72/2020\|EPI_ISL_652338\|20 | ···················· | ···················· | ····················· | ···················· | ···················· | ···················· |
| S | 2313 hCoV-19/England/NORT-286D90/2020\|EPI_ISL_652339\|20 | ···················· | ···················· | ····················· | ···················· | ···················· | ···················· |
| S | 2314 hCoV-19/England/NORT-286E33/2020\|EPI_ISL_652340\|20 | ···················· | ···················· | ····················· | ···················· | ···················· | ···················· |
| S | 2315 hCoV-19/England/NORT-286EBB/2020\|EPI_ISL_652341\|20 | ···················· | ···················· | ····················· | ···················· | ···················· | ···················· |
| S | 2316 hCoV-19/England/NORT-286F6D/2020\|EPI_ISL_652342\|20 | ···················· | ···················· | ····················· | ···················· | ···················· | ···················· |
| S | 2317 hCoV-19/England/NORT-28CEA6/2020\|EPI_ISL_652343\|20 | ···················· | ···················· | ····················· | ···················· | ···················· | ···················· |
| S | 2318 hCoV-19/England/NORT-28CF3A/2020\|EPI_ISL_652344\|20 | ···················· | ···················· | ····················· | ···················· | ···················· | ···················· |
| S | 2319 hCoV-19/England/NORT-2A0FB8/2020\|EPI_ISL_652345\|20 | ···················· | ···················· | ····················· | ···················· | ···················· | ···················· |
| S | 2320 hCoV-19/England/NORT-2A0FF4/2020\|EPI_ISL_652346\|20 | ···················· | ···················· | ····················· | ···················· | ···················· | ···················· |

| | | | | | | | |
|---|---|---|---|---|---|---|---|
| S | 2321 hCoV-19/England/NORT-2A102D/2020\|EPI_ISL_652347\|20 | ···················· | ···················· | ···················· | ···················· | ···················· | ···················· |
| S | 2323 hCoV-19/England/NORW-F2DEC/2020\|EPI_ISL_652349\|202 | ···················· | ···················· | ···················· | ···················· | ···················· | ···················· |
| S | 2324 hCoV-19/England/NORW-F610A/2020\|EPI_ISL_652350\|202 | ···················· | ···················· | ···················· | ···················· | ···················· | ···················· |
| S | 2325 hCoV-19/England/NORW-F6920/2020\|EPI_ISL_652351\|202 | ···················· | ···················· | ···················· | ···················· | ···················· | ···················· |
| S | 2327 hCoV-19/England/NORW-F6A5A/2020\|EPI_ISL_652353\|202 | ···················· | ···················· | ···················· | ···················· | ···················· | ···················· |
| S | 2329 hCoV-19/England/NORW-F6AB4/2020\|EPI_ISL_652355\|202 | ···················· | ·A·················· | ···················· | ···················· | ···················· | ··A················· |
| S | 2331 hCoV-19/England/NORW-F6B2A/2020\|EPI_ISL_652357\|202 | ···················· | ·A·················· | ···················· | ···················· | ···················· | ··A················· |
| S | 2332 hCoV-19/England/NORW-F6BFD/2020\|EPI_ISL_652358\|202 | ···················· | ···················· | ···················· | ···················· | ···················· | ···················· |
| S | 2336 hCoV-19/England/NORW-F7E99/2020\|EPI_ISL_652362\|202 | ···················· | ···················· | ···················· | ···················· | ···················· | ···················· |
| S | 2337 hCoV-19/England/NORW-F7EE4/2020\|EPI_ISL_652363\|202 | ···················· | ···················· | ···················· | ···················· | ···················· | ···················· |
| S | 2338 hCoV-19/England/NOTT-11AE08/2020\|EPI_ISL_652364\|20 | ···················· | ···················· | ···················· | ···················· | ···················· | ···················· |
| S | 2339 hCoV-19/England/NOTT-11B432/2020\|EPI_ISL_652365\|20 | ···················· | ···················· | ···················· | ···················· | ···················· | ···················· |
| S | 2340 hCoV-19/England/NOTT-11B441/2020\|EPI_ISL_652366\|20 | ···················· | ···················· | ···················· | ···················· | ···················· | ···················· |
| S | 2341 hCoV-19/England/NOTT-11B47E/2020\|EPI_ISL_652367\|20 | ···················· | ···················· | ···················· | ···················· | ···················· | ···················· |
| S | 2342 hCoV-19/England/NOTT-11B48D/2020\|EPI_ISL_652368\|20 | ···················· | ···················· | ···················· | ···················· | ···················· | ···················· |
| S | 2343 hCoV-19/England/NOTT-11B5B7/2020\|EPI_ISL_652369\|20 | ···················· | ···················· | ···················· | ···················· | ···················· | ···················· |
| S | 2344 hCoV-19/England/NOTT-11B6C3/2020\|EPI_ISL_652370\|20 | ···················· | ···················· | ···················· | ···················· | ···················· | ···················· |
| S | 2345 hCoV-19/England/NOTT-11B6D2/2020\|EPI_ISL_652371\|20 | ···················· | ···················· | ···················· | ···················· | ···················· | ···················· |
| S | 2346 hCoV-19/England/NOTT-11B6E1/2020\|EPI_ISL_652372\|20 | ···················· | ···················· | ···················· | ···················· | ···················· | ···················· |
| S | 2347 hCoV-19/England/NOTT-11B72A/2020\|EPI_ISL_652373\|20 | ···················· | ···················· | ···················· | ···················· | ···················· | ···················· |
| S | 2348 hCoV-19/England/NOTT-11B739/2020\|EPI_ISL_652374\|20 | ···················· | ···················· | ···················· | ···················· | ···················· | ···················· |
| S | 2349 hCoV-19/England/NOTT-11B757/2020\|EPI_ISL_652375\|20 | ···················· | ···················· | ···················· | ···················· | ···················· | ···················· |
| S | 2350 hCoV-19/England/NOTT-11B766/2020\|EPI_ISL_652376\|20 | ···················· | ···················· | ···················· | ···················· | ···················· | ···················· |
| S | 2351 hCoV-19/England/NOTT-11B7A2/2020\|EPI_ISL_652377\|20 | ···················· | ···················· | ···················· | ···················· | ···················· | ···················· |
| S | 2352 hCoV-19/England/NOTT-11B7B1/2020\|EPI_ISL_652378\|20 | ···················· | ···················· | ···················· | ···················· | ···················· | ···················· |
| S | 2353 hCoV-19/England/NOTT-11B7DF/2020\|EPI_ISL_652379\|20 | ···················· | ···················· | ···················· | ···················· | ···················· | ···················· |
| S | 2354 hCoV-19/England/NOTT-11BAA9/2020\|EPI_ISL_652380\|20 | ···················· | ···················· | ···················· | ···················· | ···················· | ···················· |
| S | 2357 hCoV-19/England/NOTT-11BC49/2020\|EPI_ISL_652383\|20 | ···················· | ···················· | ···················· | ···················· | ···················· | ···················· |
| S | 2358 hCoV-19/England/NOTT-11BD28/2020\|EPI_ISL_652384\|20 | ···················· | ···················· | ···················· | ···················· | ···················· | ···················· |
| S | 2359 hCoV-19/England/NOTT-11BD46/2020\|EPI_ISL_652385\|20 | ···················· | ···················· | ···················· | ···················· | ···················· | ···················· |
| S | 2360 hCoV-19/England/NOTT-11BD73/2020\|EPI_ISL_652386\|20 | ···················· | ···················· | ···················· | ···················· | ···················· | ···················· |
| S | 2361 hCoV-19/England/NOTT-11BDA0/2020\|EPI_ISL_652387\|20 | ···················· | ···················· | ···················· | ···················· | ···················· | ···················· |
| S | 2362 hCoV-19/England/NOTT-11BE70/2020\|EPI_ISL_652388\|20 | ···················· | ···················· | ···················· | ···················· | ···················· | ···················· |
| S | 2363 hCoV-19/England/NOTT-11BEAD/2020\|EPI_ISL_652389\|20 | ···················· | ···················· | ···················· | ···················· | ···················· | ···················· |
| S | 2364 hCoV-19/England/NOTT-11BEBC/2020\|EPI_ISL_652390\|20 | ···················· | ···················· | ···················· | ···················· | ···················· | ···················· |
| S | 2366 hCoV-19/England/NOTT-11BEDA/2020\|EPI_ISL_652392\|20 | ···················· | ···················· | ···················· | ···················· | ···················· | ···················· |
| S | 2367 hCoV-19/England/NOTT-11BEE9/2020\|EPI_ISL_652393\|20 | ···················· | ···················· | ···················· | ···················· | ···················· | ···················· |
| S | 2368 hCoV-19/England/NOTT-11BFD7/2020\|EPI_ISL_652394\|20 | ···················· | ···················· | ···················· | ···················· | ···················· | ···················· |
| S | 2369 hCoV-19/England/NOTT-11C04C/2020\|EPI_ISL_652395\|20 | ···················· | ···················· | ···················· | ···················· | ···················· | ···················· |
| S | 2370 hCoV-19/England/NOTT-11C079/2020\|EPI_ISL_652396\|20 | ···················· | ···················· | ···················· | ···················· | ···················· | ···················· |
| S | 2371 hCoV-19/England/NOTT-11C088/2020\|EPI_ISL_652397\|20 | ···················· | ···················· | ···················· | ···················· | ···················· | ···················· |
| S | 2372 hCoV-19/England/NOTT-11C11C/2020\|EPI_ISL_652398\|20 | ···················· | ···················· | ···················· | ···················· | ···················· | ···················· |
| S | 2373 hCoV-19/England/NOTT-11C149/2020\|EPI_ISL_652399\|20 | ···················· | ···················· | ···················· | ···················· | ···················· | ···················· |
| S | 2374 hCoV-19/England/NOTT-11C38F/2020\|EPI_ISL_652405\|20 | ···················· | ···················· | ···················· | ···················· | ···················· | ···················· |
| S | 2375 hCoV-19/England/NOTT-11C5A7/2020\|EPI_ISL_652406\|20 | ···················· | ···················· | ···················· | ···················· | ···················· | ···················· |
| S | 2376 hCoV-19/England/NOTT-11C71A/2020\|EPI_ISL_652408\|20 | ···················· | ···················· | ···················· | ···················· | ···················· | ···················· |
| S | 2377 hCoV-19/England/NOTT-11C747/2020\|EPI_ISL_652409\|20 | ···················· | ···················· | ···················· | ···················· | ···················· | ···················· |
| S | 2378 hCoV-19/USA/LA-EVTL975/2020\|EPI_ISL_653666\|2020-08 | ···················· | ···················· | ···················· | ···················· | ···················· | ···················· |
| S | 2379 hCoV-19/USA/LA-EVTL976/2020\|EPI_ISL_653667\|2020-08 | ···················· | ···················· | ···················· | ···················· | ···················· | ···················· |
| S | 2380 hCoV-19/USA/LA-EVTL977/2020\|EPI_ISL_653668\|2020-08 | ···················· | ···················· | ···················· | ···················· | ···················· | ···················· |
| S | 2381 hCoV-19/USA/LA-EVTL978/2020\|EPI_ISL_653669\|2020-08 | ···················· | ···················· | ···················· | ···················· | ···················· | ···················· |
| S | 2382 hCoV-19/USA/LA-EVTL979/2020\|EPI_ISL_653670\|2020-08 | ···················· | ···················· | ···················· | ···················· | ···················· | ···················· |
| S | 2383 hCoV-19/USA/LA-EVTL980/2020\|EPI_ISL_653671\|2020-08 | ···················· | ···················· | ···················· | ···················· | ···················· | ···················· |
| S | 2384 hCoV-19/USA/LA-EVTL981/2020\|EPI_ISL_653672\|2020-08 | ···················· | ···················· | ···················· | ···················· | ···················· | ···················· |
| S | 2385 hCoV-19/USA/LA-EVTL982/2020\|EPI_ISL_653673\|2020-08 | ···················· | ···················· | ···················· | ···················· | ···················· | ···················· |
| S | 2386 hCoV-19/USA/LA-EVTL983/2020\|EPI_ISL_653674\|2020-08 | ···················· | ···················· | ···················· | ···················· | ···················· | ···················· |
| S | 2387 hCoV-19/USA/LA-EVTL984/2020\|EPI_ISL_653675\|2020-08 | ···················· | ···················· | ···················· | ···················· | ···················· | ···················· |
| S | 2388 hCoV-19/USA/LA-EVTL985/2020\|EPI_ISL_653676\|2020-08 | ···················· | ···················· | ···················· | ···················· | ···················· | ···················· |
| S | 2389 hCoV-19/USA/LA-EVTL986/2020\|EPI_ISL_653677\|2020-08 | ···················· | ···················· | ···················· | ···················· | ···················· | ···················· |
| S | 2390 hCoV-19/USA/LA-EVTL987/2020\|EPI_ISL_653678\|2020-08 | ···················· | ···················· | ···················· | ···················· | ···················· | ···················· |
| S | 2391 hCoV-19/USA/LA-EVTL988/2020\|EPI_ISL_653679\|2020-08 | ···················· | ·A·················· | ···················· | ···················· | ···················· | ···················· |
| S | 2392 hCoV-19/USA/LA-EVTL989/2020\|EPI_ISL_653680\|2020-08 | ···················· | ···················· | ···················· | ···················· | ···················· | ···················· |
| S | 2393 hCoV-19/USA/LA-EVTL990/2020\|EPI_ISL_653681\|2020-08 | ···················· | ·A·················· | ···················· | ···················· | ···················· | ···················· |
| S | 2394 hCoV-19/USA/LA-EVTL991/2020\|EPI_ISL_653682\|2020-08 | ···················· | ···················· | ···················· | ···················· | ···················· | ···················· |
| S | 2395 hCoV-19/USA/LA-EVTL992/2020\|EPI_ISL_653683\|2020-08 | ···················· | ···················· | ···················· | ···················· | ···················· | ···················· |
| S | 2396 hCoV-19/USA/LA-EVTL993/2020\|EPI_ISL_653684\|2020-08 | ···················· | ···················· | ···················· | ···················· | ···················· | ···················· |
| S | 2397 hCoV-19/USA/LA-EVTL994/2020\|EPI_ISL_653685\|2020-08 | ···················· | ···················· | ···················· | ···················· | ···················· | ···················· |
| S | 2398 hCoV-19/USA/LA-EVTL995/2020\|EPI_ISL_653686\|2020-08 | ···················· | ···················· | ···················· | ···················· | ···················· | ···················· |
| S | 2399 hCoV-19/USA/LA-EVTL996/2020\|EPI_ISL_653687\|2020-08 | ···················· | ···················· | ···················· | ···················· | ···················· | ···················· |
| S | 2400 hCoV-19/USA/LA-EVTL997/2020\|EPI_ISL_653688\|2020-08 | ···················· | ·A·················· | ···················· | ···················· | ···················· | ···················· |
| S | 2401 hCoV-19/USA/LA-EVTL998/2020\|EPI_ISL_653689\|2020-08 | ···················· | ···················· | ···················· | ···················· | ···················· | ···················· |
| S | 2402 hCoV-19/USA/LA-EVTL999/2020\|EPI_ISL_653690\|2020-08 | ···················· | ···················· | ···················· | ···················· | ···················· | ···················· |
| S | 2403 hCoV-19/USA/LA-EVTL1000/2020\|EPI_ISL_653691\|2020-0 | ···················· | ···················· | ···················· | ···················· | ···················· | ···················· |
| S | 2404 hCoV-19/USA/LA-EVTL1001/2020\|EPI_ISL_653692\|2020-0 | ···················· | ···················· | ···················· | ···················· | ···················· | ···················· |
| S | 2405 hCoV-19/USA/LA-EVTL1002/2020\|EPI_ISL_653693\|2020-0 | ···················· | ···················· | ···················· | ···················· | ···················· | ···················· |
| S | 2406 hCoV-19/USA/LA-EVTL1003/2020\|EPI_ISL_653694\|2020-0 | ···················· | ···················· | ···················· | ···················· | ···················· | ···················· |
| S | 2407 hCoV-19/USA/LA-EVTL1004/2020\|EPI_ISL_653695\|2020-0 | ···················· | ···················· | ···················· | ···················· | ···················· | ···················· |
| S | 2408 hCoV-19/USA/LA-EVTL1005/2020\|EPI_ISL_653696\|2020-0 | ···················· | ···················· | ···················· | ···················· | ···················· | ···················· |
| S | 2409 hCoV-19/USA/LA-EVTL1006/2020\|EPI_ISL_653697\|2020-0 | ···················· | ···················· | ···················· | ···················· | ···················· | ···················· |
| S | 2410 hCoV-19/USA/LA-EVTL1007/2020\|EPI_ISL_653698\|2020-0 | ···················· | ···················· | ···················· | ···················· | ···················· | ···················· |

| | | | | | | | |
|---|---|---|---|---|---|---|---|
| S | 2411 hCoV-19/USA/LA-EVTL1008/2020\|EPI_ISL_653699\|2020-0 | ···················· | ···················· | ···················· | ···················· | ···················· | ···················· |
| S | 2412 hCoV-19/USA/LA-EVTL1009/2020\|EPI_ISL_653700\|2020-0 | ···················· | ···················· | ···················· | ···················· | ···················· | ···················· |
| S | 2413 hCoV-19/USA/LA-EVTL1010/2020\|EPI_ISL_653701\|2020-0 | ···················· | ···················· | ···················· | ···················· | ···················· | ···················· |
| S | 2414 hCoV-19/USA/LA-EVTL1011/2020\|EPI_ISL_653702\|2020-0 | ···················· | ···················· | ···················· | ···················· | ···················· | ···················· |
| S | 2415 hCoV-19/USA/LA-EVTL1012/2020\|EPI_ISL_653703\|2020-0 | ···················· | ···················· | ···················· | ···················· | ···················· | ···················· |
| S | 2416 hCoV-19/USA/LA-EVTL1013/2020\|EPI_ISL_653704\|2020-0 | ···················· | ···················· | ···················· | ···················· | ···················· | ···················· |
| S | 2417 hCoV-19/USA/LA-EVTL1014/2020\|EPI_ISL_653705\|2020-0 | ···················· | ···················· | ···················· | ···················· | ···················· | ···················· |
| S | 2418 hCoV-19/USA/LA-EVTL1015/2020\|EPI_ISL_653706\|2020-0 | ···················· | ···················· | ···················· | ···················· | ···················· | ···················· |
| S | 2419 hCoV-19/USA/LA-EVTL1016/2020\|EPI_ISL_653707\|2020-0 | ···················· | ···················· | ···················· | ···················· | ···················· | ···················· |
| S | 2420 hCoV-19/USA/LA-EVTL1017/2020\|EPI_ISL_653708\|2020-0 | ···················· | ···················· | ···················· | ···················· | ···················· | ···················· |
| S | 2421 hCoV-19/USA/LA-EVTL1018/2020\|EPI_ISL_653709\|2020-0 | ···················· | ·A·················· | ···················· | ···················· | ···················· | ···················· |
| S | 2422 hCoV-19/USA/LA-EVTL1019/2020\|EPI_ISL_653710\|2020-0 | ···················· | ·A·················· | ···················· | ···················· | ···················· | ···················· |
| S | 2423 hCoV-19/USA/LA-EVTL1020/2020\|EPI_ISL_653711\|2020-0 | ···················· | ·A·················· | ···················· | ···················· | ···················· | ···················· |
| S | 2424 hCoV-19/USA/LA-EVTL1021/2020\|EPI_ISL_653712\|2020-0 | ···················· | ·A·················· | ···················· | ···················· | ···················· | ···················· |
| S | 2425 hCoV-19/USA/LA-EVTL1022/2020\|EPI_ISL_653713\|2020-0 | ···················· | ···················· | ···················· | ···················· | ···················· | ···················· |
| S | 2426 hCoV-19/USA/LA-EVTL1023/2020\|EPI_ISL_653714\|2020-0 | ···················· | ···················· | ···················· | ···················· | ···················· | ···················· |
| S | 2427 hCoV-19/USA/LA-EVTL1024/2020\|EPI_ISL_653715\|2020-0 | ···················· | ···················· | ···················· | ···················· | ···················· | ···················· |
| S | 2428 hCoV-19/USA/LA-EVTL1025/2020\|EPI_ISL_653716\|2020-0 | ···················· | ···················· | ···················· | ···················· | ···················· | ···················· |
| S | 2429 hCoV-19/USA/LA-EVTL1026/2020\|EPI_ISL_653717\|2020-0 | ···················· | ···················· | ···················· | ···················· | ···················· | ···················· |
| S | 2430 hCoV-19/USA/LA-EVTL1027/2020\|EPI_ISL_653718\|2020-0 | ···················· | ···················· | ···················· | ···················· | ···················· | ···················· |
| S | 2431 hCoV-19/USA/LA-EVTL1028/2020\|EPI_ISL_653719\|2020-0 | ···················· | ···················· | ···················· | ···················· | ···················· | ···················· |
| S | 2432 hCoV-19/USA/LA-EVTL1029/2020\|EPI_ISL_653720\|2020-0 | ···················· | ···················· | ···················· | ···················· | ···················· | ···················· |
| S | 2433 hCoV-19/USA/LA-EVTL1046/2020\|EPI_ISL_653736\|2020-0 | ···················· | ···················· | ···················· | ···················· | ···················· | ···················· |
| S | 2434 hCoV-19/USA/LA-EVTL1047/2020\|EPI_ISL_653737\|2020-0 | ···················· | ···················· | ···················· | ···················· | ···················· | ···················· |
| S | 2435 hCoV-19/England/QEUH-B6BA0A/2020\|EPI_ISL_654997\|20 | ···················· | ···················· | ···················· | ···················· | ···················· | ···················· |
| S | 2436 hCoV-19/England/QEUH-B669C6/2020\|EPI_ISL_654998\|20 | ···················· | ···················· | ···················· | ···················· | ·····T·············· | ···················· |
| S | 2437 hCoV-19/England/QEUH-B68584/2020\|EPI_ISL_654999\|20 | ····G··············· | ···················· | ···················· | ···················· | ···················· | ···················· |
| S | 2438 hCoV-19/England/QEUH-B4367D/2020\|EPI_ISL_657200\|20 | ···················· | ···················· | ···················· | ···················· | ···················· | ···················· |
| S | 2439 hCoV-19/England/QEUH-B42562/2020\|EPI_ISL_657204\|20 | ···················· | ···················· | ···················· | ···················· | ···················· | ···················· |
| S | 2440 hCoV-19/England/QEUH-B40607/2020\|EPI_ISL_657205\|20 | ···················· | ···················· | ···················· | ···················· | ···················· | ···················· |
| S | 2441 hCoV-19/England/QEUH-B42517/2020\|EPI_ISL_657206\|20 | ···················· | ···················· | ···················· | ···················· | ···················· | ···················· |
| S | 2442 hCoV-19/England/QEUH-B40555/2020\|EPI_ISL_657207\|20 | ···················· | ···················· | ···················· | ···················· | ···················· | ···················· |
| S | 2443 hCoV-19/England/QEUH-B400FA/2020\|EPI_ISL_657208\|20 | ···················· | ···················· | ···················· | ···················· | ···················· | ···················· |
| S | 2444 hCoV-19/England/QEUH-B425BD/2020\|EPI_ISL_657209\|20 | ···················· | ···················· | ···················· | ···················· | ···················· | ···················· |
| S | 2445 hCoV-19/England/QEUH-B40670/2020\|EPI_ISL_657211\|20 | ···················· | ···················· | ···················· | ···················· | ···················· | ···················· |
| S | 2446 hCoV-19/England/QEUH-B4331C/2020\|EPI_ISL_657212\|20 | ···················· | ···················· | ···················· | ···················· | ···················· | ···················· |
| S | 2447 hCoV-19/England/QEUH-B42614/2020\|EPI_ISL_657213\|20 | ···················· | ···················· | ···················· | ···················· | ···················· | ···················· |
| S | 2448 hCoV-19/England/QEUH-B4231D/2020\|EPI_ISL_657215\|20 | ···················· | ···················· | ···················· | ···················· | ···················· | ···················· |
| S | 2449 hCoV-19/England/QEUH-B42605/2020\|EPI_ISL_657217\|20 | ···················· | ·A·················· | ··A················· | ···················· | ···················· | ··A················· |
| S | 2450 hCoV-19/England/QEUH-B405A0/2020\|EPI_ISL_657218\|20 | ···················· | ···················· | ···················· | ···················· | ···················· | ···················· |
| S | 2451 hCoV-19/England/QEUH-B423FF/2020\|EPI_ISL_657219\|20 | ···················· | ···················· | ···················· | ···················· | ···················· | ···················· |
| S | 2452 hCoV-19/England/QEUH-B40027/2020\|EPI_ISL_657220\|20 | ···················· | ···················· | ···················· | ···················· | ···················· | ···················· |
| S | 2453 hCoV-19/England/QEUH-B425AE/2020\|EPI_ISL_657222\|20 | ···················· | ···················· | ···················· | ···················· | ···················· | ···················· |
| S | 2454 hCoV-19/England/QEUH-B3FEC7/2020\|EPI_ISL_657223\|20 | ···················· | ···················· | ···················· | ···················· | ···················· | ···················· |
| S | 2455 hCoV-19/England/QEUH-B4266F/2020\|EPI_ISL_657224\|20 | ···················· | ···················· | ···················· | ···················· | ···················· | ···················· |
| S | 2456 hCoV-19/England/QEUH-B409A4/2020\|EPI_ISL_657225\|20 | ···················· | ···················· | ···················· | ···················· | ···················· | ···················· |
| S | 2457 hCoV-19/England/QEUH-B43376/2020\|EPI_ISL_657226\|20 | ···················· | ···················· | ···················· | ···················· | ···················· | ···················· |
| S | 2458 hCoV-19/England/QEUH-B40203/2020\|EPI_ISL_657227\|20 | ···················· | ···················· | ···················· | ···················· | ···················· | ···················· |
| S | 2459 hCoV-19/England/QEUH-B437C5/2020\|EPI_ISL_657228\|20 | ···················· | ·A·················· | ···················· | ···················· | ···················· | ··A················· |
| S | 2460 hCoV-19/England/QEUH-B40722/2020\|EPI_ISL_657229\|20 | ···················· | ···················· | ···················· | ···················· | ···················· | ···················· |
| S | 2461 hCoV-19/England/QEUH-B3F5B3/2020\|EPI_ISL_658517\|20 | ···················· | ···················· | ···················· | ···················· | ···················· | ···················· |
| S | 2462 hCoV-19/England/QEUH-B3F7BD/2020\|EPI_ISL_658519\|20 | ···················· | ···················· | ···················· | ···················· | ···················· | ···················· |
| S | 2463 hCoV-19/England/QEUH-B4190D/2020\|EPI_ISL_658520\|20 | ···················· | ·A·················· | ···················· | ···················· | ···················· | ···················· |
| S | 2464 hCoV-19/England/QEUH-B41A0A/2020\|EPI_ISL_658521\|20 | ···················· | ···················· | ···················· | ···················· | ···················· | ···················· |
| S | 2465 hCoV-19/England/QEUH-B416AC/2020\|EPI_ISL_658522\|20 | ···················· | ···················· | ···················· | ···················· | ···················· | ···················· |
| S | 2466 hCoV-19/England/QEUH-B3FD8E/2020\|EPI_ISL_658523\|20 | ···················· | ···················· | ···················· | ···················· | ···················· | ···················· |
| S | 2467 hCoV-19/England/QEUH-B3F3D7/2020\|EPI_ISL_658525\|20 | ···················· | ···················· | ···················· | ···················· | ···················· | ···················· |
| S | 2468 hCoV-19/England/QEUH-B3F841/2020\|EPI_ISL_658526\|20 | ···················· | ···················· | ···················· | ···················· | ···················· | ···················· |
| S | 2469 hCoV-19/England/QEUH-B3F498/2020\|EPI_ISL_658527\|20 | ···················· | ···················· | ···················· | ···················· | ···················· | ···················· |
| S | 2470 hCoV-19/England/QEUH-B41AA0/2020\|EPI_ISL_658530\|20 | ···················· | ···················· | ···················· | ···················· | ···················· | ···················· |
| S | 2471 hCoV-19/England/QEUH-B3FC72/2020\|EPI_ISL_658535\|20 | ···················· | ···················· | ···················· | ···················· | ···················· | ···················· |
| S | 2472 hCoV-19/England/QEUH-B41527/2020\|EPI_ISL_658536\|20 | ···················· | ···················· | ···················· | ···················· | ···················· | ···················· |
| S | 2473 hCoV-19/England/QEUH-B3F96C/2020\|EPI_ISL_658537\|20 | ···················· | ···················· | ···················· | ···················· | ···················· | ···················· |
| S | 2474 hCoV-19/England/QEUH-B3F665/2020\|EPI_ISL_658538\|20 | ···················· | ···················· | ···················· | ···················· | ·····T·············· | ···················· |
| S | 2475 hCoV-19/England/QEUH-B3F568/2020\|EPI_ISL_658540\|20 | ···················· | ···················· | ···················· | ···················· | ···················· | ···················· |
| S | 2476 hCoV-19/England/QEUH-B41387/2020\|EPI_ISL_658541\|20 | ···················· | ···················· | ···················· | ···················· | ···················· | ···················· |
| S | 2477 hCoV-19/England/QEUH-B415AF/2020\|EPI_ISL_658542\|20 | ···················· | ···················· | ···················· | ···················· | ···················· | ···················· |
| S | 2478 hCoV-19/England/QEUH-B3F36E/2020\|EPI_ISL_658543\|20 | ···················· | ···················· | ···················· | ···················· | ···················· | ···················· |
| S | 2479 hCoV-19/England/QEUH-B41888/2020\|EPI_ISL_658544\|20 | ···················· | ···················· | ···················· | ···················· | ···················· | ···················· |
| S | 2480 hCoV-19/England/QEUH-B41299/2020\|EPI_ISL_658545\|20 | ···················· | ···················· | ···················· | ···················· | ···················· | ···················· |
| S | 2481 hCoV-19/England/QEUH-B3F50E/2020\|EPI_ISL_658546\|20 | ···················· | ···················· | ···················· | ···················· | ···················· | ···················· |
| S | 2482 hCoV-19/England/QEUH-B4128A/2020\|EPI_ISL_658547\|20 | ···················· | ···················· | ···················· | ···················· | ···················· | ···················· |
| S | 2483 hCoV-19/England/QEUH-B3F8E7/2020\|EPI_ISL_658548\|20 | ···················· | ···················· | ···················· | ···················· | ···················· | ···················· |
| S | 2484 hCoV-19/England/QEUH-B3FC54/2020\|EPI_ISL_658549\|20 | ···················· | ···················· | ···················· | ···················· | ···················· | ···················· |
| S | 2485 hCoV-19/England/QEUH-B41A28/2020\|EPI_ISL_658550\|20 | ···················· | ···················· | ···················· | ···················· | ···················· | ···················· |
| S | 2486 hCoV-19/England/QEUH-B414DF/2020\|EPI_ISL_658551\|20 | ···················· | ···················· | ···················· | ···················· | ···················· | ···················· |
| S | 2487 hCoV-19/England/QEUH-B41ADD/2020\|EPI_ISL_658552\|20 | ···················· | ·A·················· | ···················· | ···················· | ···················· | ···················· |
| S | 2488 hCoV-19/England/QEUH-B418B5/2020\|EPI_ISL_658553\|20 | ···················· | ···················· | ···················· | ···················· | ···················· | ···················· |
| S | 2489 hCoV-19/England/QEUH-B41633/2020\|EPI_ISL_658554\|20 | ···················· | ···················· | ···················· | ···················· | ···················· | ···················· |
| S | 2490 hCoV-19/England/QEUH-B41949/2020\|EPI_ISL_658555\|20 | ···················· | ···················· | ···················· | ···················· | ···················· | ···················· |

| | | | | | | | |
|---|---|---|---|---|---|---|---|
| S | 2491 hCoV-19/England/QEUH-B3F46B/2020\|EPI_ISL_658556\|20 | ···················· | ···················· | ···················· | ···················· | ···················· | ···················· |
| S | 2492 hCoV-19/England/QEUH-B410BD/2020\|EPI_ISL_658557\|20 | ···················· | ···················· | ···················· | ···················· | ···················· | ···················· |
| S | 2493 hCoV-19/England/QEUH-B3F42F/2020\|EPI_ISL_658558\|20 | ···················· | ···················· | ···················· | ···················· | ···················· | ···················· |
| S | 2494 hCoV-19/England/QEUH-B410CC/2020\|EPI_ISL_658559\|20 | ···················· | ···················· | ···················· | ···················· | ···················· | ··A················· |
| S | 2495 hCoV-19/England/QEUH-B2F404/2020\|EPI_ISL_659800\|20 | ···················· | ···················· | ···················· | ···················· | ···················· | ···················· |
| S | 2496 hCoV-19/England/QEUH-B2F63B/2020\|EPI_ISL_659801\|20 | ···················· | ···················· | ···················· | ···················· | ···················· | ···················· |
| S | 2497 hCoV-19/England/QEUH-B2CDFD/2020\|EPI_ISL_659802\|20 | ···················· | ···················· | ···················· | ···················· | ···················· | ···················· |
| S | 2498 hCoV-19/England/QEUH-B2E8DC/2020\|EPI_ISL_659803\|20 | ···················· | ···················· | ···················· | ···················· | ···················· | ···················· |
| S | 2499 hCoV-19/England/QEUH-B2EECB/2020\|EPI_ISL_659849\|20 | ···················· | ···················· | ···················· | ···················· | ···················· | ···················· |
| S | 2500 hCoV-19/England/QEUH-B2F941/2020\|EPI_ISL_659851\|20 | ···················· | ···················· | ···················· | ···················· | ···················· | ···················· |
| S | 2501 hCoV-19/England/QEUH-B3038B/2020\|EPI_ISL_659852\|20 | ···················· | ·A··G··············· | ···················· | ···················· | ···················· | ···················· |
| S | 2502 hCoV-19/England/QEUH-B30303/2020\|EPI_ISL_659853\|20 | ···················· | ···················· | ···················· | ···················· | ···················· | ···················· |
| S | 2503 hCoV-19/England/QEUH-B2E9D9/2020\|EPI_ISL_659854\|20 | ···················· | ···················· | ···················· | ···················· | ···················· | ···················· |
| S | 2504 hCoV-19/England/QEUH-B2E942/2020\|EPI_ISL_659855\|20 | ···················· | ···················· | ···················· | ···················· | ···················· | ···················· |
| S | 2505 hCoV-19/England/QEUH-B2E001/2020\|EPI_ISL_659856\|20 | ···················· | ···················· | ···················· | ···················· | ···················· | ···················· |
| S | 2506 hCoV-19/England/QEUH-B2DF32/2020\|EPI_ISL_659857\|20 | ···················· | ···················· | ···················· | ···················· | ···················· | ···················· |
| S | 2507 hCoV-19/England/QEUH-B2F5C5/2020\|EPI_ISL_659858\|20 | ···················· | ···················· | ···················· | ···················· | ···················· | ···················· |
| S | 2508 hCoV-19/England/QEUH-B2F307/2020\|EPI_ISL_659861\|20 | ···················· | ···················· | ···················· | ···················· | ···················· | ···················· |
| S | 2509 hCoV-19/England/QEUH-B2D8EC/2020\|EPI_ISL_659866\|20 | ···················· | ···················· | ···················· | ···················· | ···················· | ···················· |
| S | 2510 hCoV-19/England/QEUH-B2D3BE/2020\|EPI_ISL_659867\|20 | ···················· | ···················· | ···················· | ···················· | ···················· | ···················· |
| S | 2511 hCoV-19/England/QEUH-B2EC49/2020\|EPI_ISL_659868\|20 | ···················· | ···················· | ···················· | ···················· | ···················· | ···················· |
| S | 2512 hCoV-19/England/QEUH-B2F686/2020\|EPI_ISL_659869\|20 | ···················· | ·A·················· | ···················· | ···················· | ···················· | ···················· |
| S | 2513 hCoV-19/England/QEUH-B2EC1C/2020\|EPI_ISL_659871\|20 | ···················· | ···················· | ···················· | ···················· | ···················· | ···················· |
| S | 2514 hCoV-19/England/QEUH-B2CE09/2020\|EPI_ISL_659872\|20 | ···················· | ···················· | ···················· | ···················· | ···················· | ···················· |
| S | 2515 hCoV-19/England/QEUH-B2CD84/2020\|EPI_ISL_659873\|20 | ···················· | ···················· | ···················· | ···················· | ···················· | ···················· |
| S | 2516 hCoV-19/England/QEUH-B3045B/2020\|EPI_ISL_659874\|20 | ···················· | ···················· | ···················· | ···················· | ···················· | ···················· |
| S | 2517 hCoV-19/England/QEUH-B2D284/2020\|EPI_ISL_659875\|20 | ···················· | ···················· | ···················· | ···················· | ···················· | ···················· |
| S | 2518 hCoV-19/England/QEUH-B2F361/2020\|EPI_ISL_659876\|20 | ···················· | ···················· | ···················· | ···················· | ···················· | ···················· |
| S | 2519 hCoV-19/England/QEUH-B2EAE5/2020\|EPI_ISL_659877\|20 | ···················· | ···················· | ···················· | ···················· | ···················· | ···················· |
| S | 2520 hCoV-19/England/QEUH-B2EE34/2020\|EPI_ISL_659878\|20 | ···················· | ·A·················· | ···················· | ···················· | ···················· | ···················· |
| S | 2521 hCoV-19/England/QEUH-B303C7/2020\|EPI_ISL_659879\|20 | ···················· | ···················· | ···················· | ···················· | ···················· | ···················· |
| S | 2522 hCoV-19/England/QEUH-B2D390/2020\|EPI_ISL_659880\|20 | ···················· | ···················· | ···················· | ···················· | ···················· | ···················· |
| S | 2523 hCoV-19/England/QEUH-B2CD39/2020\|EPI_ISL_659881\|20 | ···················· | ···················· | ···················· | ···················· | ···················· | ···················· |
| S | 2524 hCoV-19/England/QEUH-B3044C/2020\|EPI_ISL_659882\|20 | ···················· | ···················· | ···················· | ···················· | ···················· | ···················· |
| S | 2525 hCoV-19/England/QEUH-B2D6D3/2020\|EPI_ISL_659883\|20 | ···················· | ···················· | ···················· | ···················· | ···················· | ···················· |
| S | 2526 hCoV-19/England/QEUH-B2CEFA/2020\|EPI_ISL_659884\|20 | ···················· | ···················· | ···················· | ···················· | ······T············· | ···················· |
| S | 2527 hCoV-19/England/QEUH-B2E9CA/2020\|EPI_ISL_659886\|20 | ···················· | ···················· | ···················· | ···················· | ···················· | ···················· |
| S | 2528 hCoV-19/England/QEUH-B2EE25/2020\|EPI_ISL_659887\|20 | ···················· | ···················· | ···················· | ···················· | ···················· | ···················· |
| S | 2529 hCoV-19/England/QEUH-B2D925/2020\|EPI_ISL_659889\|20 | ···················· | ···················· | ···················· | ···················· | ···················· | ···················· |
| S | 2530 hCoV-19/USA/WI-GMF-21221/2020\|EPI_ISL_660830\|2020- | ···················· | ···················· | ···················· | ···················· | ···················· | ···················· |
| S | 2531 hCoV-19/USA/IA-GMF-21344/2020\|EPI_ISL_660831\|2020- | ···················· | ·············A······ | ···················· | ···················· | ···················· | ···················· |
| S | 2532 hCoV-19/USA/MN-GMF-21384/2020\|EPI_ISL_660832\|2020- | ···················· | ···················· | ···················· | ···················· | ···················· | ···················· |
| S | 2533 hCoV-19/USA/WI-GMF-21425/2020\|EPI_ISL_660833\|2020- | ···················· | ···················· | ···················· | ···················· | ···················· | ···················· |
| S | 2534 hCoV-19/USA/WI-GMF-21444/2020\|EPI_ISL_660834\|2020- | ···················· | ···················· | ···················· | ···················· | ···················· | ···············A···· |
| S | 2535 hCoV-19/USA/WI-GMF-21570/2020\|EPI_ISL_660835\|2020- | ···················· | ···················· | ···················· | ···················· | ···················· | ···················· |
| S | 2536 hCoV-19/USA/WI-GMF-21571/2020\|EPI_ISL_660836\|2020- | ···················· | ···················· | ···················· | ···················· | ···················· | ···················· |
| S | 2537 hCoV-19/USA/WI-GMF-21701/2020\|EPI_ISL_660837\|2020- | ···················· | ···················· | ···················· | ···················· | ···················· | ···················· |
| S | 2538 hCoV-19/USA/WI-GMF-21746/2020\|EPI_ISL_660838\|2020- | ···················· | ···················· | ···················· | ···················· | ···················· | ···················· |
| S | 2539 hCoV-19/USA/WI-GMF-21773/2020\|EPI_ISL_660839\|2020- | ···················· | ···················· | ···················· | ···················· | ···················· | ···············A···· |
| S | 2540 hCoV-19/USA/WI-GMF-22104/2020\|EPI_ISL_660850\|2020- | ···················· | ···················· | ···················· | ···················· | ···················· | ···················· |
| S | 2541 hCoV-19/USA/WI-GMF-22117/2020\|EPI_ISL_660851\|2020- | ···················· | ···················· | ···················· | ···················· | ···················· | ···················· |
| S | 2542 hCoV-19/USA/WI-GMF-22170/2020\|EPI_ISL_660852\|2020- | ···················· | ···················· | ···················· | ···················· | ···················· | ···················· |
| S | 2543 hCoV-19/USA/WI-GMF-22242/2020\|EPI_ISL_660853\|2020- | ···················· | ···················· | ···················· | ···················· | ···················· | ···················· |
| S | 2544 hCoV-19/USA/WI-GMF-22325/2020\|EPI_ISL_660854\|2020- | ···················· | ···················· | ···················· | ···················· | ···················· | ···················· |
| S | 2545 hCoV-19/USA/WI-GMF-22343/2020\|EPI_ISL_660855\|2020- | ···················· | ···················· | ···················· | ···················· | ···················· | ···················· |
| S | 2546 hCoV-19/USA/MN-GMF-22344/2020\|EPI_ISL_660856\|2020- | ···················· | ···················· | ···················· | ···················· | ···················· | ···················· |
| S | 2547 hCoV-19/USA/WI-GMF-22466/2020\|EPI_ISL_660857\|2020- | ···················· | ···················· | ···················· | ···················· | ···················· | ···················· |
| S | 2548 hCoV-19/USA/WI-GMF-22611/2020\|EPI_ISL_660858\|2020- | ···················· | ···················· | ···················· | ···················· | ···················· | ···················· |
| S | 2549 hCoV-19/USA/WI-GMF-22765/2020\|EPI_ISL_660859\|2020- | ···················· | ···················· | ···················· | ···················· | ···················· | ···················· |
| S | 2550 hCoV-19/USA/MN-GMF-22820/2020\|EPI_ISL_660860\|2020- | ···················· | ···················· | ···················· | ···················· | ···················· | ···················· |
| S | 2551 hCoV-19/USA/WI-GMF-22828/2020\|EPI_ISL_660861\|2020- | ···················· | ···················· | ···················· | ···················· | ···················· | ···················· |
| S | 2552 hCoV-19/USA/WI-GMF-22853/2020\|EPI_ISL_660862\|2020- | ···················· | ···················· | ···················· | ···················· | ···················· | ···················· |
| S | 2553 hCoV-19/USA/WI-GMF-22906/2020\|EPI_ISL_660863\|2020- | ···················· | ···················· | ···················· | ···················· | ···················· | ···················· |
| S | 2554 hCoV-19/USA/WI-GMF-22921/2020\|EPI_ISL_660864\|2020- | ···················· | ···················· | ···················· | ···················· | ···················· | ···················· |
| S | 2555 hCoV-19/USA/WI-GMF-22933/2020\|EPI_ISL_660865\|2020- | ···················· | ···················· | ···················· | ···················· | ···················· | ···················· |
| S | 2556 hCoV-19/USA/WI-GMF-23113/2020\|EPI_ISL_660866\|2020- | ···················· | ···················· | ···················· | ···················· | ···················· | ···················· |
| S | 2557 hCoV-19/USA/WI-GMF-23209/2020\|EPI_ISL_660867\|2020- | ···················· | ···················· | ···················· | ···················· | ···················· | ···················· |
| S | 2558 hCoV-19/USA/WI-GMF-23240/2020\|EPI_ISL_660868\|2020- | ···················· | ···················· | ···················· | ···················· | ···················· | ···············A···· |
| S | 2559 hCoV-19/USA/WI-GMF-23259/2020\|EPI_ISL_660869\|2020- | ···················· | ···················· | ···················· | ···················· | ···················· | ···················· |
| S | 2560 hCoV-19/USA/WI-GMF-23445/2020\|EPI_ISL_660870\|2020- | ···················· | ···················· | ···················· | ···················· | ···················· | ···················· |
| S | 2561 hCoV-19/England/NORW-F6823/2020\|EPI_ISL_651090\|202 | ···················· | ···················· | ···················· | ···················· | ···················· | ···················· |
| S | 2562 hCoV-19/England/NORW-F687E/2020\|EPI_ISL_651091\|202 | ···················· | ···················· | ···················· | ···················· | ···················· | ···················· |
| S | 2563 hCoV-19/England/NORW-F6B48/2020\|EPI_ISL_651092\|202 | ···················· | ···················· | ···················· | ···················· | ···················· | ···················· |
| S | 2565 hCoV-19/England/NORW-F7F1E/2020\|EPI_ISL_651094\|202 | ···················· | ···················· | ···················· | ···················· | ···················· | ···················· |
| S | 2566 hCoV-19/England/LIVE-DB4271/2020\|EPI_ISL_651095\|20 | ···················· | ···················· | ···················· | ···················· | ···················· | ···················· |
| S | 2567 hCoV-19/England/MILK-B055C5/2020\|EPI_ISL_651096\|20 | ···················· | ···················· | ···················· | ···················· | ···················· | ···················· |
| S | 2568 hCoV-19/Northern Ireland/NIRE-1091A9/2020\|EPI_ISL_ | ···················· | ···················· | ···················· | ···················· | ···················· | ···················· |
| S | 2569 hCoV-19/England/BIRM-6AA15/2020\|EPI_ISL_651098\|202 | ···················· | ·A·················· | ···················· | ···················· | ···················· | ··A················· |
| S | 2570 hCoV-19/England/MILK-A23FD3/2020\|EPI_ISL_651099\|20 | ···················· | ·A·················· | ···················· | ···················· | ···················· | ··A················· |
| S | 2571 hCoV-19/England/QEUH-B42508/2020\|EPI_ISL_657110\|20 | ···················· | ·A·················· | ··A················· | ···················· | ···················· | ··A················· |

S 2572 hCoV-19/England/QEUH-B43983/2020|EPI_ISL_657111|20 ···················· ···················· ····················· ···················· ···················· ····················
S 2573 hCoV-19/England/QEUH-B40528/2020|EPI_ISL_657112|20 ···················· ···················· ····················· ···················· ···················· ····················
S 2574 hCoV-19/England/QEUH-B427D5/2020|EPI_ISL_657113|20 ···················· ···················· ····················· ···················· ···················· ····················
S 2575 hCoV-19/England/QEUH-B4222F/2020|EPI_ISL_657117|20 ···················· ···················· ····················· ···················· ···················· ····················
S 2576 hCoV-19/England/QEUH-B434FB/2020|EPI_ISL_657118|20 ···················· ···················· ····················· ···················· ···················· ····················
S 2577 hCoV-19/England/QEUH-B42395/2020|EPI_ISL_657119|20 ···················· ···················· ····················· ···················· ···················· ····················
S 2578 hCoV-19/England/QEUH-B3FED6/2020|EPI_ISL_657130|20 ···················· ···················· ····················· ···················· ···················· ····················
S 2579 hCoV-19/England/QEUH-B43534/2020|EPI_ISL_657131|20 ···················· ···················· ····················· ···················· ······T············· ····················
S 2580 hCoV-19/England/QEUH-B43349/2020|EPI_ISL_657132|20 ···················· ···················· ····················· ···················· ···················· ····················
S 2581 hCoV-19/England/QEUH-B40643/2020|EPI_ISL_657133|20 ···················· ···················· ····················· ···················· ···················· ····················
S 2582 hCoV-19/England/QEUH-B40573/2020|EPI_ISL_657134|20 ···················· ···················· ····················· ···················· ···················· ····················
S 2583 hCoV-19/England/QEUH-B3FE21/2020|EPI_ISL_657135|20 ···················· ···················· ····················· ···················· ···················· ····················
S 2584 hCoV-19/England/QEUH-B408B6/2020|EPI_ISL_657136|20 ···················· ···················· ····················· ···················· ···················· ····················
S 2585 hCoV-19/England/QEUH-B40467/2020|EPI_ISL_657137|20 ···················· ·A·················· ····················· ···················· ···················· ···A················
S 2586 hCoV-19/England/QEUH-B43491/2020|EPI_ISL_657138|20 ···················· ···················· ····················· ···················· ···················· ····················
S 2587 hCoV-19/England/QEUH-B433A3/2020|EPI_ISL_657139|20 ···················· ·A·················· ····················· ···················· ···················· ···A················
S 2588 hCoV-19/England/QEUH-B4374D/2020|EPI_ISL_657140|20 ···················· ···················· ····················· ···················· ···················· ····················
S 2589 hCoV-19/England/QEUH-B3FF00/2020|EPI_ISL_657141|20 ···················· ···················· ····················· ···················· ···················· ····················
S 2590 hCoV-19/England/QEUH-B42104/2020|EPI_ISL_657142|20 ···················· ···················· ····················· ···················· ···················· ····················
S 2591 hCoV-19/England/QEUH-B436AA/2020|EPI_ISL_657143|20 ···················· ···················· ····················· ···················· ···················· ····················
S 2592 hCoV-19/England/QEUH-B40063/2020|EPI_ISL_657144|20 ···················· ···················· ····················· ···················· ···················· ····················
S 2593 hCoV-19/England/QEUH-B40889/2020|EPI_ISL_657145|20 ···················· ···················· ····················· ···················· ···················· ····················
S 2594 hCoV-19/England/QEUH-B42799/2020|EPI_ISL_657146|20 ···················· ···················· ····················· ···················· ···················· ····················
S 2595 hCoV-19/England/QEUH-B42359/2020|EPI_ISL_657147|20 ···················· ···················· ····················· ···················· ···················· ····················
S 2596 hCoV-19/England/QEUH-B43455/2020|EPI_ISL_657148|20 ···················· ···················· ····················· ···················· ···················· ····················
S 2597 hCoV-19/England/QEUH-B424ED/2020|EPI_ISL_657150|20 ···················· ···················· ····················· ···················· ···················· ····················
S 2598 hCoV-19/England/QEUH-B3FDE8/2020|EPI_ISL_657151|20 ···················· ···················· ····················· ···················· ···················· ····················
S 2599 hCoV-19/England/QEUH-B40081/2020|EPI_ISL_657152|20 ···················· ···················· ····················· ···················· ···················· ····················
S 2600 hCoV-19/England/QEUH-B4233B/2020|EPI_ISL_657153|20 ···················· ···················· ····················· ···················· ···················· ····················
S 2601 hCoV-19/England/QEUH-B40106/2020|EPI_ISL_657154|20 ···················· ···················· ····················· ···················· ···················· ····················
S 2602 hCoV-19/England/QEUH-B40731/2020|EPI_ISL_657155|20 ···················· ···················· ····················· ···················· ···················· ····················
S 2603 hCoV-19/England/QEUH-B404FE/2020|EPI_ISL_657157|20 ···················· ···················· ····················· ···················· ···················· ····················
S 2604 hCoV-19/England/QEUH-B40212/2020|EPI_ISL_657158|20 ···················· ·A·················· ····················· ···················· ···················· ···A················
S 2605 hCoV-19/England/QEUH-B437A7/2020|EPI_ISL_657159|20 ···················· ···················· ····················· ···················· ···················· ····················
S 2606 hCoV-19/England/QEUH-B405EC/2020|EPI_ISL_657160|20 ···················· ·A·················· ····················· ···················· ···················· ····················
S 2607 hCoV-19/England/QEUH-B40142/2020|EPI_ISL_657161|20 ···················· ·A·················· ····················· ···················· ···················· ···A················
S 2608 hCoV-19/England/QEUH-B3FE7C/2020|EPI_ISL_657162|20 ···················· ···················· ····················· ···················· ···················· ····················
S 2609 hCoV-19/England/QEUH-B4087A/2020|EPI_ISL_657163|20 ···················· ···················· ····················· ···················· ···················· ····················
S 2610 hCoV-19/England/QEUH-B40704/2020|EPI_ISL_657164|20 ···················· ···················· ····················· ···················· ···················· ····················
S 2611 hCoV-19/England/QEUH-B425CC/2020|EPI_ISL_657165|20 ···················· ···················· ····················· ···················· ···················· ····················
S 2612 hCoV-19/England/QEUH-B436E6/2020|EPI_ISL_657166|20 ···················· ···················· ····················· ···················· ···················· ····················
S 2613 hCoV-19/England/QEUH-B406DA/2020|EPI_ISL_657167|20 ···················· ···················· ····················· ···················· ···················· ····················
S 2614 hCoV-19/England/QEUH-B3FE9A/2020|EPI_ISL_657168|20 ···················· ···················· ····················· ···················· ···················· ····················
S 2615 hCoV-19/England/QEUH-B423A4/2020|EPI_ISL_657169|20 ···················· ···················· ····················· ···················· ···················· ····················
S 2616 hCoV-19/England/QEUH-B42289/2020|EPI_ISL_657170|20 ···················· ···················· ····················· ···················· ···················· ····················
S 2617 hCoV-19/England/QEUH-B407F5/2020|EPI_ISL_657171|20 ···················· ···················· ····················· ···················· ···················· ····················
S 2618 hCoV-19/England/QEUH-B4225C/2020|EPI_ISL_657172|20 ···················· ···················· ····················· ···················· ···················· ····················
S 2619 hCoV-19/England/QEUH-B4024F/2020|EPI_ISL_657173|20 ···················· ·A·················· ····················· ···················· ···················· ···A················
S 2620 hCoV-19/England/QEUH-B40625/2020|EPI_ISL_657174|20 ···················· ···················· ····················· ···················· ···················· ····················
S 2621 hCoV-19/England/QEUH-B407AA/2020|EPI_ISL_657175|20 ···················· ···················· ····················· ···················· ···················· ····················
S 2622 hCoV-19/England/QEUH-B40652/2020|EPI_ISL_657176|20 ···················· ···················· ····················· ···················· ···················· ····················
S 2623 hCoV-19/England/QEUH-B4091D/2020|EPI_ISL_657177|20 ···················· ···················· ····················· ···················· ···················· ····················
S 2624 hCoV-19/England/QEUH-B3FE6D/2020|EPI_ISL_657178|20 ···················· ···················· ····················· ···················· ···················· ····················
S 2625 hCoV-19/England/QEUH-B40009/2020|EPI_ISL_657179|20 ···················· ···················· ····················· ···················· ···················· ····················
S 2626 hCoV-19/England/QEUH-B42544/2020|EPI_ISL_657180|20 ···················· ·A·················· ···A················· ···················· ···················· ···A················
S 2627 hCoV-19/England/QEUH-B421E6/2020|EPI_ISL_657181|20 ···················· ···················· ····················· ···················· ···················· ····················
S 2628 hCoV-19/England/QEUH-B433C1/2020|EPI_ISL_657182|20 ···················· ·A·················· ····················· ···················· ···················· ···A················
S 2629 hCoV-19/England/QEUH-B4050A/2020|EPI_ISL_657183|20 ···················· ···················· ····················· ···················· ···················· ····················
S 2630 hCoV-19/England/QEUH-B4219B/2020|EPI_ISL_657184|20 ···················· ···················· ····················· ···················· ···················· ····················
S 2631 hCoV-19/England/QEUH-B405CE/2020|EPI_ISL_657185|20 ···················· ···················· ····················· ···················· ···················· ····················
S 2632 hCoV-19/England/QEUH-B425DB/2020|EPI_ISL_657186|20 ···················· ···················· ····················· ···················· ···················· ····················
S 2633 hCoV-19/England/QEUH-B40160/2020|EPI_ISL_657187|20 ···················· ···················· ····················· ···················· ···················· ····················
S 2634 hCoV-19/England/QEUH-B3FF2E/2020|EPI_ISL_657188|20 ···················· ···················· ····················· ···················· ···················· ····················
S 2635 hCoV-19/England/QEUH-B4079B/2020|EPI_ISL_657189|20 ···················· ···················· ····················· ···················· ···················· ····················
S 2636 hCoV-19/England/QEUH-B420F8/2020|EPI_ISL_657201|20 ···················· ···················· ····················· ···················· ···················· ····················
S 2637 hCoV-19/England/QEUH-B43428/2020|EPI_ISL_657203|20 ···················· ···················· ····················· ···················· ···················· ····················
S 2638 hCoV-19/England/QEUH-B3F8D8/2020|EPI_ISL_658440|20 ···················· ···················· ····················· ···················· ···················· ····················
S 2639 hCoV-19/England/QEUH-B3F4F2/2020|EPI_ISL_658441|20 ···················· ···················· ····················· ···················· ···················· ····················
S 2640 hCoV-19/England/QEUH-B416F7/2020|EPI_ISL_658442|20 ···················· ···················· ····················· ···················· ···················· ····················
S 2641 hCoV-19/England/QEUH-B3F86F/2020|EPI_ISL_658443|20 ···················· ···················· ····················· ···················· ···················· ····················
S 2642 hCoV-19/England/QEUH-B3F4C5/2020|EPI_ISL_658444|20 ···················· ···················· ····················· ···················· ···················· ····················
S 2643 hCoV-19/England/QEUH-B3F313/2020|EPI_ISL_658447|20 ···················· ···················· ····················· ···················· ···················· ····················
S 2644 hCoV-19/England/QEUH-B3F225/2020|EPI_ISL_658448|20 ···················· ···················· ····················· ···················· ···················· ····················
S 2645 hCoV-19/England/QEUH-B3F3F5/2020|EPI_ISL_658449|20 ···················· ···················· ····················· ···················· ···················· ····················
S 2646 hCoV-19/England/QEUH-B3F89C/2020|EPI_ISL_658460|20 ···················· ···················· ····················· ···················· ···················· ····················
S 2647 hCoV-19/England/QEUH-B3FA0F/2020|EPI_ISL_658461|20 ···················· ···················· ····················· ···················· ···················· ····················
S 2648 hCoV-19/England/QEUH-B414EE/2020|EPI_ISL_658462|20 ···················· ·········T·········· ····················· ···················· ···················· ····················
S 2649 hCoV-19/England/QEUH-B41466/2020|EPI_ISL_658463|20 ···················· ···················· ····················· ···················· ···················· ····················
S 2650 hCoV-19/England/QEUH-B3F3C8/2020|EPI_ISL_658464|20 ···················· ···················· ····················· ···················· ···················· ····················
S 2651 hCoV-19/England/QEUH-B3FCFA/2020|EPI_ISL_658465|20 ···················· ···················· ····················· ···················· ···················· ····················

| | | | | | | | |
|---|---|---|---|---|---|---|---|
| S | 2652 hCoV-19/England/QEUH-B3FA3C/2020\|EPI_ISL_658466\|20 | ···················· | ···················· | ····················· | ···················· | ···················· | ···············R···· |
| S | 2653 hCoV-19/England/QEUH-B416E8/2020\|EPI_ISL_658467\|20 | ···················· | ···················· | ····················· | ···················· | ···················· | ···················· |
| S | 2654 hCoV-19/England/QEUH-B40F84/2020\|EPI_ISL_658468\|20 | ···················· | ···················· | ····················· | ···················· | ···················· | ···················· |
| S | 2655 hCoV-19/England/QEUH-B3F410/2020\|EPI_ISL_658469\|20 | ···················· | ···················· | ····················· | ···················· | ···················· | ···················· |
| S | 2656 hCoV-19/England/QEUH-B3FC36/2020\|EPI_ISL_658470\|20 | ···················· | ···················· | ····················· | ···················· | ···················· | ···················· |
| S | 2657 hCoV-19/England/QEUH-B4182E/2020\|EPI_ISL_658471\|20 | ···················· | ···················· | ····················· | ···················· | ···················· | ···················· |
| S | 2658 hCoV-19/England/QEUH-B3F29E/2020\|EPI_ISL_658472\|20 | ···················· | ···················· | ····················· | ···················· | ···················· | ···················· |
| S | 2659 hCoV-19/England/QEUH-B41A64/2020\|EPI_ISL_658473\|20 | ···················· | ···················· | ····················· | ···················· | ···················· | ···················· |
| S | 2660 hCoV-19/England/QEUH-B3F920/2020\|EPI_ISL_658474\|20 | ···················· | ···················· | ····················· | ···················· | ···················· | ···················· |
| S | 2661 hCoV-19/England/QEUH-B410F9/2020\|EPI_ISL_658475\|20 | ···················· | ···················· | ····················· | ···················· | ···················· | ···················· |
| S | 2662 hCoV-19/England/QEUH-B3FB2A/2020\|EPI_ISL_658476\|20 | ···················· | ···················· | ····················· | ···················· | ···················· | ···················· |
| S | 2663 hCoV-19/England/QEUH-B3FD06/2020\|EPI_ISL_658477\|20 | ···················· | ···················· | ····················· | ···················· | ···················· | ···················· |
| S | 2664 hCoV-19/England/QEUH-B3F3B9/2020\|EPI_ISL_658478\|20 | ···················· | ···················· | ····················· | ···················· | ···················· | ···················· |
| S | 2665 hCoV-19/England/QEUH-B3F9E4/2020\|EPI_ISL_658479\|20 | ···················· | ···················· | ····················· | ···················· | ···················· | ···················· |
| S | 2666 hCoV-19/England/QEUH-B3F43E/2020\|EPI_ISL_658480\|20 | ···················· | ·A·················· | ····················· | ···················· | ···················· | ··A················· |
| S | 2667 hCoV-19/England/QEUH-B4123F/2020\|EPI_ISL_658481\|20 | ···················· | ·A·················· | ····················· | ···················· | ···················· | ··A················· |
| S | 2668 hCoV-19/England/QEUH-B4168E/2020\|EPI_ISL_658482\|20 | ···················· | ···················· | ····················· | ···················· | ···················· | ···················· |
| S | 2669 hCoV-19/England/QEUH-B3F6B0/2020\|EPI_ISL_658483\|20 | ···················· | ···················· | ····················· | ···················· | ···················· | ···················· |
| S | 2670 hCoV-19/England/QEUH-B4142A/2020\|EPI_ISL_658484\|20 | ···················· | ···················· | ····················· | ···················· | ···················· | ···················· |
| S | 2671 hCoV-19/England/QEUH-B40F48/2020\|EPI_ISL_658485\|20 | ···················· | ···················· | ····················· | ···················· | ···················· | ···················· |
| S | 2672 hCoV-19/England/QEUH-B417D6/2020\|EPI_ISL_658486\|20 | ···················· | ···················· | ····················· | ···················· | ···················· | ···················· |
| S | 2673 hCoV-19/England/QEUH-B3F780/2020\|EPI_ISL_658487\|20 | ···················· | ···················· | ····················· | ···················· | ···················· | ···················· |
| S | 2674 hCoV-19/England/QEUH-B414B1/2020\|EPI_ISL_658488\|20 | ···················· | ···················· | ····················· | ···················· | ···················· | ···················· |
| S | 2675 hCoV-19/England/QEUH-B4175E/2020\|EPI_ISL_658491\|20 | ···················· | ···················· | ····················· | ···················· | ···················· | ···················· |
| S | 2676 hCoV-19/England/QEUH-B3F7EA/2020\|EPI_ISL_658492\|20 | ···················· | ···················· | ····················· | ···················· | ···················· | ···················· |
| S | 2677 hCoV-19/England/QEUH-B412B7/2020\|EPI_ISL_658493\|20 | ···················· | ···················· | ····················· | ···················· | ···················· | ···················· |
| S | 2678 hCoV-19/England/QEUH-B3F331/2020\|EPI_ISL_658495\|20 | ···················· | ···················· | ····················· | ···················· | ···················· | ···················· |
| S | 2679 hCoV-19/England/QEUH-B3F9F3/2020\|EPI_ISL_658496\|20 | ···················· | ···················· | ····················· | ···················· | ···················· | ···················· |
| S | 2680 hCoV-19/England/QEUH-B41080/2020\|EPI_ISL_658497\|20 | ···················· | ···················· | ····················· | ···················· | ···················· | ···················· |
| S | 2681 hCoV-19/England/QEUH-B413B4/2020\|EPI_ISL_658498\|20 | ···················· | ···················· | ····················· | ···················· | ···················· | ···················· |
| S | 2682 hCoV-19/England/QEUH-B3F4A7/2020\|EPI_ISL_658499\|20 | ···················· | ···················· | ····················· | ···················· | ···················· | ···················· |
| S | 2683 hCoV-19/England/QEUH-B412A8/2020\|EPI_ISL_658500\|20 | ···················· | ···················· | ····················· | ···················· | ···················· | ···················· |
| S | 2684 hCoV-19/England/QEUH-B3F8C9/2020\|EPI_ISL_658501\|20 | ···················· | ···················· | ····················· | ···················· | ···················· | ···················· |
| S | 2685 hCoV-19/England/QEUH-B414A2/2020\|EPI_ISL_658502\|20 | ···················· | ···················· | ····················· | ···················· | ···················· | ···················· |
| S | 2686 hCoV-19/England/QEUH-B417B8/2020\|EPI_ISL_658503\|20 | ···················· | ···················· | ····················· | ···················· | ···················· | ···················· |
| S | 2687 hCoV-19/England/QEUH-B3FAB4/2020\|EPI_ISL_658508\|20 | ···················· | ···················· | ····················· | ···················· | ···················· | ···················· |
| S | 2688 hCoV-19/England/QEUH-B3F35F/2020\|EPI_ISL_658509\|20 | ···················· | ···················· | ····················· | ···················· | ···················· | ···················· |
| S | 2689 hCoV-19/England/QEUH-B41484/2020\|EPI_ISL_658510\|20 | ···················· | ···················· | ····················· | ···················· | ···················· | ···················· |
| S | 2690 hCoV-19/England/QEUH-B4186A/2020\|EPI_ISL_658511\|20 | ···················· | ···················· | ····················· | ···················· | ···················· | ···················· |
| S | 2691 hCoV-19/England/QEUH-B3F577/2020\|EPI_ISL_658513\|20 | ···················· | ·A·················· | ····················· | ···················· | ···················· | ···················· |
| S | 2692 hCoV-19/England/QEUH-B4191C/2020\|EPI_ISL_658514\|20 | ···················· | ·A·················· | ····················· | ···················· | ···················· | ···················· |
| S | 2693 hCoV-19/England/QEUH-B41150/2020\|EPI_ISL_658515\|20 | ···················· | ···················· | ····················· | ···················· | ···················· | ···················· |
| S | 2694 hCoV-19/England/QEUH-B3F52C/2020\|EPI_ISL_658516\|20 | ···················· | ···················· | ····················· | ···················· | ···················· | ···················· |
| S | 2695 hCoV-19/England/QEUH-B417A9/2020\|EPI_ISL_658528\|20 | ···················· | ···················· | ····················· | ···················· | ···················· | ···················· |
| S | 2696 hCoV-19/England/QEUH-B41B07/2020\|EPI_ISL_658529\|20 | ···················· | ···················· | ····················· | ···················· | ···················· | ···················· |
| S | 2697 hCoV-19/England/QEUH-B3FAE1/2020\|EPI_ISL_658531\|20 | ···················· | ···················· | ····················· | ···················· | ···················· | ···················· |
| S | 2698 hCoV-19/England/QEUH-B3F3E6/2020\|EPI_ISL_658533\|20 | ···················· | ···················· | ····················· | ···················· | ···················· | ···················· |
| S | 2699 hCoV-19/England/QEUH-B41730/2020\|EPI_ISL_658534\|20 | ···················· | ···················· | ····················· | ···················· | ···················· | ···················· |
| S | 2700 hCoV-19/England/QEUH-B2EC3A/2020\|EPI_ISL_659770\|20 | ···················· | ···················· | ····················· | ···················· | ···················· | ···················· |
| S | 2701 hCoV-19/England/QEUH-B2D257/2020\|EPI_ISL_659771\|20 | ···················· | ···················· | ····················· | ···················· | ···················· | ···················· |
| S | 2702 hCoV-19/England/QEUH-B30AA4/2020\|EPI_ISL_659773\|20 | ···················· | ···················· | ····················· | ···················· | ···················· | ···················· |
| S | 2703 hCoV-19/England/QEUH-B2E8FA/2020\|EPI_ISL_659774\|20 | ···················· | ···················· | ····················· | ···················· | ···················· | ···················· |
| S | 2704 hCoV-19/England/QEUH-B2EF04/2020\|EPI_ISL_659775\|20 | ···················· | ···················· | ····················· | ···················· | ···················· | ···················· |
| S | 2705 hCoV-19/England/QEUH-B2EE70/2020\|EPI_ISL_659778\|20 | ···················· | ···················· | ····················· | ···················· | ···················· | ···················· |
| S | 2706 hCoV-19/England/QEUH-B2EC58/2020\|EPI_ISL_659779\|20 | ···················· | ···················· | ····················· | ···················· | ···················· | ···················· |
| S | 2707 hCoV-19/England/QEUH-B2D5C7/2020\|EPI_ISL_659790\|20 | ···················· | ···················· | ····················· | ···················· | ···················· | ···················· |
| S | 2708 hCoV-19/England/QEUH-B2DA5F/2020\|EPI_ISL_659791\|20 | ···················· | ···················· | ····················· | ···················· | ···················· | ···················· |
| S | 2709 hCoV-19/England/QEUH-B2D59A/2020\|EPI_ISL_659792\|20 | ···················· | ···················· | ····················· | ···················· | ···················· | ···················· |
| S | 2710 hCoV-19/England/QEUH-B2CF15/2020\|EPI_ISL_659793\|20 | ···················· | ···················· | ····················· | ···················· | ······T············· | ···················· |
| S | 2711 hCoV-19/England/QEUH-B2F60E/2020\|EPI_ISL_659794\|20 | ···················· | ···················· | ····················· | ···················· | ···················· | ···················· |
| S | 2712 hCoV-19/England/QEUH-B2E854/2020\|EPI_ISL_659796\|20 | ···················· | ···················· | ····················· | ···················· | ···················· | ···················· |
| S | 2713 hCoV-19/England/QEUH-B2CCB4/2020\|EPI_ISL_659797\|20 | ···················· | ···················· | ····················· | ···················· | ···················· | ···················· |
| S | 2714 hCoV-19/England/QEUH-B2EFAA/2020\|EPI_ISL_659799\|20 | ···················· | ···················· | ····················· | ···················· | ···················· | ···················· |
| S | 2715 hCoV-19/England/QEUH-B2F62C/2020\|EPI_ISL_659805\|20 | ···················· | ···················· | ····················· | ···················· | ···················· | ···················· |
| S | 2716 hCoV-19/England/QEUH-B2CC87/2020\|EPI_ISL_659806\|20 | ···················· | ···················· | ····················· | ···················· | ···················· | ···················· |
| S | 2717 hCoV-19/England/QEUH-B302F7/2020\|EPI_ISL_659807\|20 | ···················· | ···················· | ····················· | ···················· | ···················· | ···················· |
| S | 2718 hCoV-19/England/QEUH-B2D6C4/2020\|EPI_ISL_659808\|20 | ···················· | ···················· | ····················· | ···················· | ···················· | ···················· |
| S | 2719 hCoV-19/England/QEUH-B2F3DA/2020\|EPI_ISL_659810\|20 | ···················· | ···················· | ····················· | ···················· | ···················· | ···················· |
| S | 2720 hCoV-19/England/QEUH-B2F695/2020\|EPI_ISL_659811\|20 | ···················· | ···················· | ····················· | ···················· | ···················· | ···················· |
| S | 2721 hCoV-19/England/QEUH-B30B92/2020\|EPI_ISL_659813\|20 | ···················· | ···················· | ····················· | ···················· | ···················· | ···················· |
| S | 2722 hCoV-19/England/QEUH-B2EF5F/2020\|EPI_ISL_659814\|20 | ···················· | ·A·················· | ····················· | ···················· | ···················· | ···················· |
| S | 2723 hCoV-19/England/QEUH-B2D62E/2020\|EPI_ISL_659815\|20 | ···················· | ···················· | ····················· | ···················· | ···················· | ···················· |
| S | 2724 hCoV-19/England/QEUH-B2F370/2020\|EPI_ISL_659816\|20 | ···················· | ···················· | ····················· | ···················· | ···················· | ···················· |
| S | 2725 hCoV-19/England/QEUH-B30B65/2020\|EPI_ISL_659817\|20 | ···················· | ···················· | ····················· | ···················· | ···················· | ···················· |
| S | 2726 hCoV-19/England/QEUH-B2EB3D/2020\|EPI_ISL_659818\|20 | ···················· | ···················· | ····················· | ···················· | ····T··············· | ···················· |
| S | 2727 hCoV-19/England/QEUH-B2E906/2020\|EPI_ISL_659819\|20 | ···················· | ···················· | ····················· | ···················· | ···················· | ···················· |
| S | 2728 hCoV-19/England/QEUH-B3039A/2020\|EPI_ISL_659820\|20 | ···················· | ···················· | ····················· | ···················· | ···················· | ···················· |
| S | 2729 hCoV-19/England/QEUH-B2D4F7/2020\|EPI_ISL_659821\|20 | ···················· | ···················· | ····················· | ···················· | ···················· | ···················· |
| S | 2730 hCoV-19/England/QEUH-B2DA13/2020\|EPI_ISL_659822\|20 | ···················· | ···················· | ····················· | ···················· | ···················· | ···················· |
| S | 2731 hCoV-19/England/QEUH-B2D239/2020\|EPI_ISL_659823\|20 | ···················· | ···················· | ····················· | ···················· | ···················· | ···················· |

| | | | | | | | |
|---|---|---|---|---|---|---|---|
| S | 2732 hCoV-19/England/QEUH-B2D354/2020\|EPI_ISL_659825\|20 | ···················· | ···················· | ····················· | ···················· | ···················· | ···················· |
| S | 2733 hCoV-19/England/QEUH-B2D697/2020\|EPI_ISL_659827\|20 | ···················· | ···················· | ····················· | ···················· | ···················· | ···················· |
| S | 2734 hCoV-19/England/QEUH-B2D61F/2020\|EPI_ISL_659828\|20 | ···················· | ···················· | ····················· | ···················· | ···················· | ···················· |
| S | 2735 hCoV-19/England/QEUH-B2EF13/2020\|EPI_ISL_659829\|20 | ···················· | ···················· | ····················· | ···················· | ···················· | ···················· |
| S | 2736 hCoV-19/England/QEUH-B2EBD3/2020\|EPI_ISL_659830\|20 | ···················· | ···················· | ····················· | ···················· | ···················· | ···················· |
| S | 2737 hCoV-19/England/QEUH-B2E951/2020\|EPI_ISL_659831\|20 | ···················· | ···················· | ····················· | ···················· | ···················· | ···················· |
| S | 2738 hCoV-19/England/QEUH-B2DEBD/2020\|EPI_ISL_659832\|20 | ···················· | ···················· | ····················· | ···················· | ···················· | ···················· |
| S | 2739 hCoV-19/England/QEUH-B2D47F/2020\|EPI_ISL_659833\|20 | ···················· | ···················· | ····················· | ···················· | ···················· | ···················· |
| S | 2740 hCoV-19/England/QEUH-B2D266/2020\|EPI_ISL_659836\|20 | ···················· | ···················· | ····················· | ···················· | ···················· | ···················· |
| S | 2741 hCoV-19/England/QEUH-B2D530/2020\|EPI_ISL_659837\|20 | ···················· | ···················· | ····················· | ···················· | ···················· | ···················· |
| S | 2742 hCoV-19/England/QEUH-B2F89F/2020\|EPI_ISL_659838\|20 | ···················· | ···················· | ····················· | ···················· | ···················· | ···················· |
| S | 2743 hCoV-19/England/QEUH-B2DF14/2020\|EPI_ISL_659840\|20 | ···················· | ···················· | ····················· | ···················· | ···················· | ···················· |
| S | 2744 hCoV-19/England/QEUH-B2D5D6/2020\|EPI_ISL_659842\|20 | ···················· | ···················· | ····················· | ···················· | ···················· | ···················· |
| S | 2745 hCoV-19/England/QEUH-B2DF50/2020\|EPI_ISL_659843\|20 | ···················· | ···················· | ····················· | ···················· | ···················· | ···················· |
| S | 2746 hCoV-19/England/QEUH-B30AE0/2020\|EPI_ISL_659844\|20 | ···················· | ···················· | ····················· | ···················· | ···················· | ···················· |
| S | 2747 hCoV-19/England/QEUH-B2DEEA/2020\|EPI_ISL_659845\|20 | ···················· | ···················· | ····················· | ···················· | ···················· | ···················· |
| S | 2748 hCoV-19/England/QEUH-B302BB/2020\|EPI_ISL_659846\|20 | ···················· | ···················· | ····················· | ···················· | ···················· | ···················· |
| S | 2749 hCoV-19/England/QEUH-B2EDEC/2020\|EPI_ISL_659847\|20 | ···················· | ···················· | ····················· | ···················· | ···················· | ···················· |
| S | 2750 hCoV-19/England/QEUH-B2E872/2020\|EPI_ISL_659859\|20 | ···················· | ···················· | ····················· | ···················· | ···················· | ···················· |
| S | 2751 hCoV-19/England/QEUH-B2D600/2020\|EPI_ISL_659863\|20 | ···················· | ···················· | ····················· | ···················· | ···················· | ···················· |
| S | 2752 hCoV-19/England/QEUH-B2E8CD/2020\|EPI_ISL_659865\|20 | ···················· | ···················· | ····················· | ···················· | ···················· | ···················· |
| S | 2753 hCoV-19/England/204490924/2020\|EPI_ISL_660760\|2020 | ···················· | ···················· | ····················· | ···················· | ···················· | ···················· |
| S | 2754 hCoV-19/England/204491276/2020\|EPI_ISL_660772\|2020 | ···················· | ···················· | ····················· | ···················· | ···················· | ···················· |
| S | 2755 hCoV-19/England/204500124/2020\|EPI_ISL_660774\|2020 | ···················· | ···················· | ····················· | ···················· | ···················· | ···················· |
| S | 2756 hCoV-19/England/204500127/2020\|EPI_ISL_660775\|2020 | ···················· | ···················· | ····················· | ···················· | ···················· | ···················· |
| S | 2757 hCoV-19/England/204500132/2020\|EPI_ISL_660776\|2020 | ···················· | ···················· | ····················· | ···················· | ···················· | ···················· |
| S | 2758 hCoV-19/England/204500135/2020\|EPI_ISL_660777\|2020 | ···················· | ···················· | ····················· | ···················· | ···················· | ···················· |
| S | 2759 hCoV-19/England/204500136/2020\|EPI_ISL_660778\|2020 | ···················· | ···················· | ····················· | ···················· | ···················· | ···················· |
| S | 2760 hCoV-19/England/204500153/2020\|EPI_ISL_660779\|2020 | ···················· | ···················· | ····················· | ···················· | ···················· | ···················· |
| S | 2761 hCoV-19/England/204500451/2020\|EPI_ISL_660782\|2020 | ···················· | ···················· | ····················· | ···················· | ···················· | ···················· |
| S | 2762 hCoV-19/England/204500452/2020\|EPI_ISL_660783\|2020 | ···················· | ···················· | ····················· | ···················· | ···················· | ···················· |
| S | 2763 hCoV-19/England/204500453/2020\|EPI_ISL_660784\|2020 | ···················· | ···················· | ····················· | ···················· | ···················· | ···················· |
| S | 2764 hCoV-19/England/204501167/2020\|EPI_ISL_660786\|2020 | ···················· | ···················· | ····················· | ···················· | ···················· | ···················· |
| S | 2765 hCoV-19/England/204501192/2020\|EPI_ISL_660787\|2020 | ···················· | ···················· | ····················· | ···················· | ···················· | ···················· |
| S | 2766 hCoV-19/England/204501194/2020\|EPI_ISL_660788\|2020 | ···················· | ···················· | ····················· | ···················· | ···················· | ···················· |
| S | 2767 hCoV-19/England/204501201/2020\|EPI_ISL_660789\|2020 | ···················· | ···················· | ····················· | ···················· | ···················· | ···················· |
| S | 2768 hCoV-19/England/204501205/2020\|EPI_ISL_660790\|2020 | ···················· | ···················· | ····················· | ···················· | ···················· | ···················· |
| S | 2769 hCoV-19/England/204501206/2020\|EPI_ISL_660791\|2020 | ···················· | ···················· | ····················· | ···················· | ···················· | ···················· |
| S | 2770 hCoV-19/England/204521631/2020\|EPI_ISL_660792\|2020 | ···················· | ···················· | ····················· | ···················· | ···················· | ···················· |
| S | 2771 hCoV-19/England/204521640/2020\|EPI_ISL_660793\|2020 | ···················· | ···················· | ····················· | ···················· | ···················· | ···················· |
| S | 2772 hCoV-19/USA/WI-GMF-20453/2020\|EPI_ISL_660795\|2020- | ···················· | ···················· | ····················· | ···················· | ···················· | ···················· |
| S | 2773 hCoV-19/USA/WI-GMF-20481/2020\|EPI_ISL_660796\|2020- | ···················· | ···················· | ····················· | ···················· | ···················· | ···················· |
| S | 2774 hCoV-19/USA/WI-GMF-20501/2020\|EPI_ISL_660797\|2020- | ···················· | ···················· | ····················· | ···················· | ···················· | ···················· |
| S | 2775 hCoV-19/USA/WI-GMF-20504/2020\|EPI_ISL_660798\|2020- | ···················· | ···················· | ····················· | ···················· | ···················· | ···················· |
| S | 2776 hCoV-19/USA/WI-GMF-20538/2020\|EPI_ISL_660799\|2020- | ···················· | ···················· | ····················· | ···················· | ···················· | ···················· |
| S | 2777 hCoV-19/USA/WI-GMF-20543/2020\|EPI_ISL_660800\|2020- | ···················· | ···················· | ····················· | ···················· | ···················· | ···················· |
| S | 2778 hCoV-19/USA/WI-GMF-20576/2020\|EPI_ISL_660801\|2020- | ···················· | ···················· | ····················· | ···················· | ···················· | ···················· |
| S | 2779 hCoV-19/USA/WI-GMF-20586/2020\|EPI_ISL_660802\|2020- | ···················· | ···················· | ····················· | ···················· | ···················· | ···················· |
| S | 2780 hCoV-19/USA/WI-GMF-20611/2020\|EPI_ISL_660803\|2020- | ···················· | ···················· | ····················· | ···················· | ···················· | ···················· |
| S | 2781 hCoV-19/USA/WI-GMF-20612/2020\|EPI_ISL_660804\|2020- | ···················· | ···················· | ····················· | ···················· | ···················· | ···················· |
| S | 2782 hCoV-19/USA/WI-GMF-20613/2020\|EPI_ISL_660805\|2020- | ···················· | ···················· | ····················· | ···················· | ···················· | ···················· |
| S | 2783 hCoV-19/USA/WI-GMF-20617/2020\|EPI_ISL_660806\|2020- | ···················· | ···················· | ····················· | ···················· | ···················· | ···················· |
| S | 2784 hCoV-19/USA/WI-GMF-20621/2020\|EPI_ISL_660807\|2020- | ···················· | ·············A······ | ····················· | ···················· | ···················· | ···················· |
| S | 2785 hCoV-19/USA/WI-GMF-20630/2020\|EPI_ISL_660808\|2020- | ···················· | ···················· | ····················· | ···················· | ···················· | ···················· |
| S | 2786 hCoV-19/USA/WI-GMF-20643/2020\|EPI_ISL_660809\|2020- | ···················· | ···················· | ····················· | ···················· | ···················· | ···················· |
| S | 2787 hCoV-19/USA/WI-GMF-20649/2020\|EPI_ISL_660810\|2020- | ···················· | ···················· | ····················· | ···················· | ···················· | ···················· |
| S | 2788 hCoV-19/USA/WI-GMF-20712/2020\|EPI_ISL_660811\|2020- | ···················· | ···················· | ····················· | ···················· | ···················· | ···················· |
| S | 2789 hCoV-19/USA/WI-GMF-20745/2020\|EPI_ISL_660812\|2020- | ···················· | ···················· | ····················· | ···················· | ···················· | ···················· |
| S | 2790 hCoV-19/USA/WI-GMF-20760/2020\|EPI_ISL_660813\|2020- | ···················· | ···················· | ····················· | ···················· | ···················· | ···················· |
| S | 2791 hCoV-19/USA/WI-GMF-20762/2020\|EPI_ISL_660814\|2020- | ···················· | ···················· | ····················· | ···················· | ···················· | ···················· |
| S | 2792 hCoV-19/USA/WI-GMF-20782/2020\|EPI_ISL_660815\|2020- | ···················· | ···················· | ····················· | ···················· | ···················· | ···················· |
| S | 2793 hCoV-19/USA/WI-GMF-20811/2020\|EPI_ISL_660816\|2020- | ···················· | ···················· | ····················· | ···················· | ···················· | ···················· |
| S | 2794 hCoV-19/USA/WI-GMF-20840/2020\|EPI_ISL_660817\|2020- | ···················· | ···················· | ····················· | ···················· | ···················· | ···················· |
| S | 2795 hCoV-19/USA/WI-GMF-20859/2020\|EPI_ISL_660818\|2020- | ···················· | ···················· | ····················· | ···················· | ···················· | ···················· |
| S | 2796 hCoV-19/USA/WI-GMF-20861/2020\|EPI_ISL_660819\|2020- | ···················· | ···················· | ····················· | ···················· | ···················· | ···················· |
| S | 2797 hCoV-19/USA/WI-GMF-20899/2020\|EPI_ISL_660820\|2020- | ···················· | ···················· | ····················· | ···················· | ···················· | ···················· |
| S | 2798 hCoV-19/USA/WI-GMF-20912/2020\|EPI_ISL_660821\|2020- | ···················· | ···················· | ····················· | ···················· | ···················· | ···············A···· |
| S | 2799 hCoV-19/USA/WI-GMF-20969/2020\|EPI_ISL_660822\|2020- | ···················· | ···················· | ····················· | ···················· | ···················· | ···················· |
| S | 2800 hCoV-19/USA/WI-GMF-20970/2020\|EPI_ISL_660823\|2020- | ···················· | ···················· | ····················· | ···················· | ···················· | ···············A···· |
| S | 2801 hCoV-19/USA/WI-GMF-20974/2020\|EPI_ISL_660824\|2020- | ···················· | ···················· | ····················· | ···················· | ···················· | ···················· |
| S | 2802 hCoV-19/USA/MN-GMF-20997/2020\|EPI_ISL_660825\|2020- | ···················· | ···················· | ····················· | ···················· | ···················· | ···················· |
| S | 2803 hCoV-19/USA/WI-GMF-21043/2020\|EPI_ISL_660826\|2020- | ···················· | ···················· | ····················· | ···················· | ···················· | ···················· |
| S | 2804 hCoV-19/USA/WI-GMF-21045/2020\|EPI_ISL_660827\|2020- | ···················· | ···················· | ····················· | ···················· | ···················· | ···················· |
| S | 2805 hCoV-19/USA/WI-GMF-21115/2020\|EPI_ISL_660828\|2020- | ···················· | ···················· | ····················· | ···················· | ···················· | ···················· |
| S | 2806 hCoV-19/USA/IA-GMF-21220/2020\|EPI_ISL_660829\|2020- | ···················· | ···················· | ····················· | ···················· | ···················· | ···················· |
| S | 2807 hCoV-19/USA/WI-GMF-21812/2020\|EPI_ISL_660840\|2020- | ···················· | ···················· | ····················· | ···················· | ···················· | ···················· |
| S | 2808 hCoV-19/USA/MN-GMF-21836/2020\|EPI_ISL_660841\|2020- | ···················· | ···················· | ····················· | ···················· | ···················· | ···················· |
| S | 2809 hCoV-19/USA/MN-GMF-21861/2020\|EPI_ISL_660842\|2020- | ···················· | ···················· | ····················· | ···················· | ···················· | ···················· |
| S | 2810 hCoV-19/USA/WI-GMF-21955/2020\|EPI_ISL_660843\|2020- | ···················· | ···················· | ····················· | ···················· | ···················· | ···················· |
| S | 2811 hCoV-19/USA/MN-GMF-22034/2020\|EPI_ISL_660844\|2020- | ···················· | ···················· | ····················· | ···················· | ···················· | ···················· |

| | | | | | | | |
|---|---|---|---|---|---|---|---|
| S | 2812 hCoV-19/USA/WI-GMF-22035/2020\|EPI_ISL_660845\|2020- | ···················· | ···················· | ····················· | ···················· | ···················· | ···················· |
| S | 2813 hCoV-19/USA/WI-GMF-22047/2020\|EPI_ISL_660846\|2020- | ···················· | ···················· | ····················· | ···················· | ···················· | ···················· |
| S | 2814 hCoV-19/USA/WI-GMF-22070/2020\|EPI_ISL_660847\|2020- | ···················· | ···················· | ····················· | ···················· | ···················· | ···················· |
| S | 2815 hCoV-19/USA/MN-GMF-22096/2020\|EPI_ISL_660848\|2020- | ···················· | ···················· | ····················· | ···················· | ···················· | ···················· |
| S | 2816 hCoV-19/USA/WI-GMF-22102/2020\|EPI_ISL_660849\|2020- | ···················· | ···················· | ····················· | ···················· | ···················· | ···················· |
| S | 2817 hCoV-19/England/MILK-B06A98/2020\|EPI_ISL_652258\|20 | ···················· | ·A·················· | ····················· | ···················· | ···················· | ··A················· |
| S | 2818 hCoV-19/England/MILK-B06AC5/2020\|EPI_ISL_652259\|20 | ···················· | ···················· | ····················· | ···················· | ···················· | ···················· |
| S | 2819 hCoV-19/England/MILK-B06AE3/2020\|EPI_ISL_652260\|20 | ···················· | ···················· | ····················· | ···················· | ···················· | ···················· |
| S | 2820 hCoV-19/England/MILK-B06B0E/2020\|EPI_ISL_652261\|20 | ···················· | ·A·················· | ····················· | ···················· | ···················· | ··A················· |
| S | 2821 hCoV-19/England/MILK-B06B2C/2020\|EPI_ISL_652262\|20 | ···················· | ···················· | ····················· | ···················· | ···················· | ···················· |
| S | 2822 hCoV-19/England/MILK-B06B95/2020\|EPI_ISL_652263\|20 | ···················· | ·A·················· | ····················· | ···················· | ···················· | ··A················· |
| S | 2823 hCoV-19/England/MILK-B06BB3/2020\|EPI_ISL_652264\|20 | ···················· | ···················· | ····················· | ···················· | ···················· | ···················· |
| S | 2824 hCoV-19/England/MILK-B06BC2/2020\|EPI_ISL_652265\|20 | ···················· | ···················· | ····················· | ···················· | ···················· | ···················· |
| S | 2825 hCoV-19/England/MILK-B06BD1/2020\|EPI_ISL_652266\|20 | ···················· | ···················· | ····················· | ···················· | ···················· | ···················· |
| S | 2826 hCoV-19/England/MILK-B06C29/2020\|EPI_ISL_652267\|20 | ···················· | ···················· | ····················· | ···················· | ···················· | ···················· |
| S | 2827 hCoV-19/England/MILK-B06C47/2020\|EPI_ISL_652268\|20 | ···················· | ···················· | ····················· | ···················· | ···················· | ···················· |
| S | 2828 hCoV-19/England/MILK-B06C83/2020\|EPI_ISL_652270\|20 | ···················· | ···················· | ····················· | ···················· | ···················· | ···················· |
| S | 2829 hCoV-19/England/MILK-B06CB0/2020\|EPI_ISL_652271\|20 | ···················· | ···················· | ····················· | ···················· | ···················· | ···················· |
| S | 2830 hCoV-19/England/MILK-B06CDE/2020\|EPI_ISL_652272\|20 | ···················· | ·A·················· | ····················· | ···················· | ···················· | ··A················· |
| S | 2831 hCoV-19/England/MILK-B06CED/2020\|EPI_ISL_652273\|20 | ···················· | ···················· | ····················· | ···················· | ···················· | ···················· |
| S | 2832 hCoV-19/England/MILK-B06D44/2020\|EPI_ISL_652274\|20 | ···················· | ···················· | ····················· | ···················· | ···················· | ···················· |
| S | 2833 hCoV-19/England/MILK-B06D80/2020\|EPI_ISL_652275\|20 | ···················· | ···················· | ····················· | ···················· | ···················· | ···················· |
| S | 2834 hCoV-19/England/MILK-B06EAB/2020\|EPI_ISL_652280\|20 | ···················· | ···················· | ····················· | ···················· | ···················· | ···················· |
| S | 2835 hCoV-19/England/MILK-B06EC9/2020\|EPI_ISL_652281\|20 | ···················· | ···················· | ····················· | ···················· | ···················· | ···················· |
| S | 2836 hCoV-19/England/MILK-B06ED8/2020\|EPI_ISL_652282\|20 | ···················· | ···················· | ····················· | ···················· | ···················· | ···················· |
| S | 2837 hCoV-19/England/MILK-B06EF6/2020\|EPI_ISL_652283\|20 | ···················· | ···················· | ····················· | ···················· | ··········Y········· | ···················· |
| S | 2838 hCoV-19/England/MILK-B06F3F/2020\|EPI_ISL_652284\|20 | ···················· | ···················· | ····················· | ···················· | ···················· | ···················· |
| S | 2839 hCoV-19/England/MILK-B06F5D/2020\|EPI_ISL_652285\|20 | ···················· | ···················· | ····················· | ···················· | ···················· | ···················· |
| S | 2840 hCoV-19/England/MILK-B06F8A/2020\|EPI_ISL_652286\|20 | ···················· | ···················· | ····················· | ···················· | ···················· | ···················· |
| S | 2841 hCoV-19/England/MILK-B06FD5/2020\|EPI_ISL_652287\|20 | ···················· | ···················· | ····················· | ···················· | ···················· | ···················· |
| S | 2842 hCoV-19/England/MILK-B0701D/2020\|EPI_ISL_652288\|20 | ···················· | ·A·················· | ····················· | ···················· | ···················· | ··A················· |
| S | 2843 hCoV-19/England/MILK-B38E0A/2020\|EPI_ISL_652289\|20 | ···················· | ···················· | ····················· | ···················· | ···················· | ···················· |
| S | 2844 hCoV-19/England/MILK-B38F70/2020\|EPI_ISL_652290\|20 | ···················· | ···················· | ····················· | ···················· | ···················· | ···················· |
| S | 2845 hCoV-19/England/MILK-B38FE9/2020\|EPI_ISL_652291\|20 | ···················· | ···················· | ····················· | ···················· | ···················· | ···················· |
| S | 2846 hCoV-19/England/MILK-B3909A/2020\|EPI_ISL_652292\|20 | ···················· | ···················· | ····················· | ···················· | ···················· | ···················· |
| S | 2847 hCoV-19/England/MILK-B390A9/2020\|EPI_ISL_652293\|20 | ···················· | ···················· | ····················· | ···················· | ···················· | ···················· |
| S | 2848 hCoV-19/England/MILK-B3915B/2020\|EPI_ISL_652294\|20 | ···················· | ···················· | ····················· | ···················· | ···················· | ···················· |
| S | 2849 hCoV-19/England/MILK-B391D3/2020\|EPI_ISL_652295\|20 | ···················· | ···················· | ····················· | ···················· | ···················· | ···················· |
| S | 2850 hCoV-19/England/MILK-B391E2/2020\|EPI_ISL_652296\|20 | ···················· | ···················· | ····················· | ···················· | ···················· | ···················· |
| S | 2851 hCoV-19/England/MILK-B3920D/2020\|EPI_ISL_652297\|20 | ···················· | ···················· | ····················· | ···················· | ···················· | ···················· |
| S | 2852 hCoV-19/England/MILK-B3923A/2020\|EPI_ISL_652298\|20 | ···················· | ···················· | ····················· | ···················· | ···················· | ···················· |
| S | 2853 hCoV-19/England/MILK-B39294/2020\|EPI_ISL_652299\|20 | ···················· | ···················· | ····················· | ···················· | ···················· | ···················· |
| S | 2854 hCoV-19/England/MILK-B392EF/2020\|EPI_ISL_652300\|20 | ···················· | ···················· | ····················· | ···················· | ···················· | ···················· |
| S | 2855 hCoV-19/England/MILK-B39917/2020\|EPI_ISL_652301\|20 | ···················· | ···················· | ····················· | ···················· | ···················· | ···················· |
| S | 2858 hCoV-19/Northern Ireland/NIRE-108FA0/2020\|EPI_ISL_ | ···················· | ···················· | ····················· | ···················· | ···················· | ···················· |
| S | 2859 hCoV-19/Northern Ireland/NIRE-108FBF/2020\|EPI_ISL_ | ···················· | ···················· | ····················· | ···················· | ···················· | ···················· |
| S | 2860 hCoV-19/Northern Ireland/NIRE-108FCE/2020\|EPI_ISL_ | ···················· | ···················· | ····················· | ···················· | ···················· | ···················· |
| S | 2861 hCoV-19/Northern Ireland/NIRE-108FDD/2020\|EPI_ISL_ | ···················· | ···················· | ····················· | ···················· | ···················· | ···················· |
| S | 2862 hCoV-19/Northern Ireland/NIRE-108FEC/2020\|EPI_ISL_ | ···················· | ···················· | ····················· | ···················· | ···················· | ···················· |
| S | 2863 hCoV-19/Northern Ireland/NIRE-109015/2020\|EPI_ISL_ | ···················· | ···················· | ····················· | ···················· | ···················· | ···················· |
| S | 2864 hCoV-19/Northern Ireland/NIRE-109051/2020\|EPI_ISL_ | ···················· | ···················· | ····················· | ···················· | ···················· | ···················· |
| S | 2865 hCoV-19/Northern Ireland/NIRE-10909D/2020\|EPI_ISL_ | ···················· | ···················· | ····················· | ···················· | ···················· | ···················· |
| S | 2866 hCoV-19/Northern Ireland/NIRE-1090BB/2020\|EPI_ISL_ | ···················· | ···················· | ····················· | ···················· | ···················· | ···················· |
| S | 2867 hCoV-19/Northern Ireland/NIRE-1090D9/2020\|EPI_ISL_ | ···················· | ···················· | ····················· | ···················· | ···················· | ···················· |
| S | 2868 hCoV-19/Northern Ireland/NIRE-1090F7/2020\|EPI_ISL_ | ···················· | ···················· | ····················· | ···················· | ···················· | ···················· |
| S | 2869 hCoV-19/Northern Ireland/NIRE-109103/2020\|EPI_ISL_ | ···················· | ···················· | ····················· | ···················· | ···················· | ···················· |
| S | 2870 hCoV-19/Northern Ireland/NIRE-109130/2020\|EPI_ISL_ | ···················· | ···················· | ····················· | ···················· | ···················· | ···················· |
| S | 2871 hCoV-19/Northern Ireland/NIRE-10915E/2020\|EPI_ISL_ | ···················· | ···················· | ····················· | ···················· | ···················· | ···················· |
| S | 2872 hCoV-19/Northern Ireland/NIRE-10917C/2020\|EPI_ISL_ | ···················· | ···················· | ····················· | ···················· | ···················· | ···················· |
| S | 2873 hCoV-19/Northern Ireland/NIRE-10918B/2020\|EPI_ISL_ | ···················· | ···················· | ····················· | ···················· | ···················· | ···················· |
| S | 2874 hCoV-19/Northern Ireland/NIRE-1091C7/2020\|EPI_ISL_ | ···················· | ···················· | ····················· | ···················· | ···················· | ···················· |
| S | 2875 hCoV-19/Northern Ireland/NIRE-109200/2020\|EPI_ISL_ | ···················· | ···················· | ····················· | ···················· | ···················· | ···················· |
| S | 2876 hCoV-19/Northern Ireland/NIRE-10922E/2020\|EPI_ISL_ | ···················· | ···················· | ····················· | ···················· | ···················· | ···················· |
| S | 2877 hCoV-19/Northern Ireland/NIRE-10923D/2020\|EPI_ISL_ | ···················· | ···················· | ····················· | ···················· | ···················· | ···················· |
| S | 2878 hCoV-19/Northern Ireland/NIRE-10924C/2020\|EPI_ISL_ | ···················· | ···················· | ····················· | ···················· | ···················· | ···················· |
| S | 2879 hCoV-19/Northern Ireland/NIRE-109288/2020\|EPI_ISL_ | ···················· | ···················· | ····················· | ···················· | ···················· | ···················· |
| S | 2880 hCoV-19/Northern Ireland/NIRE-109297/2020\|EPI_ISL_ | ···················· | ···················· | ····················· | ···················· | ···················· | ···················· |
| S | 2881 hCoV-19/Northern Ireland/NIRE-1092D3/2020\|EPI_ISL_ | ···················· | ···················· | ····················· | ···················· | ···················· | ···················· |
| S | 2882 hCoV-19/Northern Ireland/NIRE-1092F1/2020\|EPI_ISL_ | ···················· | ···················· | ····················· | ···················· | ···················· | ···················· |
| S | 2883 hCoV-19/Northern Ireland/NIRE-10933A/2020\|EPI_ISL_ | ···················· | ···················· | ····················· | ···················· | ···················· | ···················· |
| S | 2884 hCoV-19/Northern Ireland/NIRE-109394/2020\|EPI_ISL_ | ···················· | ···················· | ····················· | ···················· | ···················· | ···················· |
| S | 2885 hCoV-19/Northern Ireland/NIRE-1093FE/2020\|EPI_ISL_ | ···················· | ···················· | ····················· | ···················· | ···················· | ···················· |
| S | 2886 hCoV-19/Northern Ireland/NIRE-1094DD/2020\|EPI_ISL_ | ···················· | ···················· | ····················· | ···················· | ···················· | ···················· |
| S | 2887 hCoV-19/Northern Ireland/NIRE-1094FB/2020\|EPI_ISL_ | ···················· | ···················· | ····················· | ···················· | ···················· | ···················· |
| S | 2888 hCoV-19/Northern Ireland/NIRE-109561/2020\|EPI_ISL_ | ···················· | ···················· | ····················· | ···················· | ···················· | ···················· |
| S | 2889 hCoV-19/USA/LA-EVTL898/2020\|EPI_ISL_653589\|2020-07 | ···················· | ···················· | ····················· | ···················· | ···················· | ···················· |
| S | 2890 hCoV-19/USA/LA-EVTL899/2020\|EPI_ISL_653590\|2020-08 | ···················· | ·············A······ | ····················· | ···················· | ···················· | ···················· |
| S | 2891 hCoV-19/USA/LA-EVTL900/2020\|EPI_ISL_653591\|2020-07 | ···················· | ···················· | ····················· | ···················· | ···················· | ···················· |
| S | 2892 hCoV-19/USA/LA-EVTL901/2020\|EPI_ISL_653592\|2020-07 | ···················· | ·A·················· | ····················· | ···················· | ···················· | ···················· |
| S | 2893 hCoV-19/USA/LA-EVTL902/2020\|EPI_ISL_653593\|2020-07 | ···················· | ···················· | ····················· | ···················· | ···················· | ···················· |

| | | | | | | | | |
|---|---|---|---|---|---|---|---|---|
| S | 2894 | hCoV-19/USA/LA-EVTL903/2020\|EPI_ISL_653594\|2020-08 | .................... | .................... | ..................... | .................... | .................... | .................... |
| S | 2895 | hCoV-19/USA/LA-EVTL904/2020\|EPI_ISL_653595\|2020-07 | .................... | .................... | ..................... | .................... | .................... | .................... |
| S | 2896 | hCoV-19/USA/LA-EVTL905/2020\|EPI_ISL_653596\|2020-07 | .................... | .................... | ..................... | .................... | .................... | .................... |
| S | 2897 | hCoV-19/USA/LA-EVTL906/2020\|EPI_ISL_653597\|2020-07 | .................... | .................... | ..................... | .................... | .................... | .................... |
| S | 2898 | hCoV-19/USA/LA-EVTL907/2020\|EPI_ISL_653598\|2020-07 | .................... | .................... | ..................... | .................... | .................... | .................... |
| S | 2899 | hCoV-19/USA/LA-EVTL908/2020\|EPI_ISL_653599\|2020-07 | .................... | .................... | ..................... | .................... | .................... | .................... |
| S | 2900 | hCoV-19/USA/LA-EVTL909/2020\|EPI_ISL_653600\|2020-08 | .................... | .................... | ..................... | .................... | .................... | .................... |
| S | 2901 | hCoV-19/USA/LA-EVTL910/2020\|EPI_ISL_653601\|2020-07 | .................... | .................... | ..................... | .................... | .................... | .................... |
| S | 2902 | hCoV-19/USA/LA-EVTL911/2020\|EPI_ISL_653602\|2020-07 | .................... | .................... | ..................... | .................... | .................... | .................... |
| S | 2903 | hCoV-19/USA/LA-EVTL912/2020\|EPI_ISL_653603\|2020-07 | .................... | .................... | ..................... | .................... | .................... | .................... |
| S | 2904 | hCoV-19/USA/LA-EVTL913/2020\|EPI_ISL_653604\|2020-07 | .................... | .................... | ..................... | .................... | .................... | .................... |
| S | 2905 | hCoV-19/USA/LA-EVTL914/2020\|EPI_ISL_653605\|2020-07 | .................... | .................... | ..................... | .................... | .................... | .................... |
| S | 2906 | hCoV-19/USA/LA-EVTL915/2020\|EPI_ISL_653606\|2020-07 | .................... | .A.................. | ..................... | .................... | .................... | .................... |
| S | 2907 | hCoV-19/USA/LA-EVTL916/2020\|EPI_ISL_653607\|2020-08 | .................... | ......A............. | ..................... | .................... | .................... | .................... |
| S | 2908 | hCoV-19/USA/LA-EVTL917/2020\|EPI_ISL_653608\|2020-07 | .................... | .................... | ..................... | .................... | .................... | .................... |
| S | 2909 | hCoV-19/USA/LA-EVTL918/2020\|EPI_ISL_653609\|2020-07 | .................... | .................... | ..................... | .................... | .................... | .................... |
| S | 2910 | hCoV-19/USA/LA-EVTL919/2020\|EPI_ISL_653610\|2020-08 | .................... | .................... | ..................... | .................... | .................... | .................... |
| S | 2911 | hCoV-19/USA/LA-EVTL920/2020\|EPI_ISL_653611\|2020-08 | .................... | .................... | ..................... | .................... | .................... | .................... |
| S | 2912 | hCoV-19/USA/LA-EVTL921/2020\|EPI_ISL_653612\|2020-08 | .................... | .................... | ..................... | .................... | .................... | .................... |
| S | 2913 | hCoV-19/USA/LA-EVTL922/2020\|EPI_ISL_653613\|2020-08 | .................... | .................... | ..................... | .................... | .................... | .................... |
| S | 2914 | hCoV-19/USA/LA-EVTL923/2020\|EPI_ISL_653614\|2020-08 | .................... | .................... | ..................... | .................... | .................... | .................... |
| S | 2915 | hCoV-19/USA/LA-EVTL924/2020\|EPI_ISL_653615\|2020-08 | .................... | .................... | ..................... | .................... | .................... | .................... |
| S | 2916 | hCoV-19/USA/LA-EVTL925/2020\|EPI_ISL_653616\|2020-08 | .................... | .................... | ..................... | .................... | .................... | .................... |
| S | 2917 | hCoV-19/USA/LA-EVTL926/2020\|EPI_ISL_653617\|2020-08 | .................... | .................... | ..................... | .................... | .................... | .................... |
| S | 2918 | hCoV-19/USA/LA-EVTL927/2020\|EPI_ISL_653618\|2020-08 | .................... | .................... | ..................... | .................... | .................... | .................... |
| S | 2919 | hCoV-19/USA/LA-EVTL928/2020\|EPI_ISL_653619\|2020-08 | .................... | .................... | ..................... | .................... | .................... | .................... |
| S | 2920 | hCoV-19/USA/LA-EVTL929/2020\|EPI_ISL_653620\|2020-08 | .................... | .................... | ..................... | .................... | .................... | .................... |
| S | 2921 | hCoV-19/USA/LA-EVTL930/2020\|EPI_ISL_653621\|2020-08 | .................... | .................... | ..................... | .................... | .................... | .................... |
| S | 2922 | hCoV-19/USA/LA-EVTL931/2020\|EPI_ISL_653622\|2020-08 | .................... | .................... | ..................... | .................... | .................... | .................... |
| S | 2923 | hCoV-19/USA/LA-EVTL932/2020\|EPI_ISL_653623\|2020-08 | .................... | .................... | ..................... | .................... | .................... | .................... |
| S | 2924 | hCoV-19/USA/LA-EVTL933/2020\|EPI_ISL_653624\|2020-08 | .................... | .................... | ..................... | .................... | .................... | .................... |
| S | 2925 | hCoV-19/USA/LA-EVTL934/2020\|EPI_ISL_653625\|2020-08 | .................... | .................... | ..................... | .................... | .................... | .................... |
| S | 2926 | hCoV-19/USA/LA-EVTL935/2020\|EPI_ISL_653626\|2020-08 | .................... | .................... | ..................... | .................... | .................... | .................... |
| S | 2927 | hCoV-19/USA/LA-EVTL936/2020\|EPI_ISL_653627\|2020-08 | .................... | .................... | ..................... | .................... | .................... | .................... |
| S | 2928 | hCoV-19/USA/LA-EVTL937/2020\|EPI_ISL_653628\|2020-08 | .................... | .................... | ..................... | .................... | .................... | .................... |
| S | 2929 | hCoV-19/USA/LA-EVTL938/2020\|EPI_ISL_653629\|2020-08 | .................... | .................... | ..................... | .................... | .................... | .................... |
| S | 2930 | hCoV-19/USA/LA-EVTL939/2020\|EPI_ISL_653630\|2020-08 | .................... | .................... | ..................... | .................... | .................... | .................... |
| S | 2931 | hCoV-19/USA/LA-EVTL940/2020\|EPI_ISL_653631\|2020-08 | .................... | .................... | ..................... | .................... | .................... | .................... |
| S | 2932 | hCoV-19/USA/LA-EVTL941/2020\|EPI_ISL_653632\|2020-08 | .................... | .................... | ..................... | .................... | .................... | .................... |
| S | 2933 | hCoV-19/USA/LA-EVTL942/2020\|EPI_ISL_653633\|2020-08 | .................... | .................... | ..................... | .................... | .................... | .................... |
| S | 2934 | hCoV-19/USA/LA-EVTL943/2020\|EPI_ISL_653634\|2020-08 | .................... | .................... | ..................... | .................... | .................... | .................... |
| S | 2935 | hCoV-19/USA/LA-EVTL944/2020\|EPI_ISL_653635\|2020-08 | .................... | .................... | ..................... | .................... | .................... | .................... |
| S | 2936 | hCoV-19/USA/LA-EVTL945/2020\|EPI_ISL_653636\|2020-08 | .................... | .................... | ..................... | .................... | .................... | .................... |
| S | 2937 | hCoV-19/USA/LA-EVTL946/2020\|EPI_ISL_653637\|2020-08 | .................... | .................... | ..................... | .................... | .................... | .................... |
| S | 2938 | hCoV-19/USA/LA-EVTL947/2020\|EPI_ISL_653638\|2020-08 | .................... | .................... | ..................... | .................... | .................... | .................... |
| S | 2939 | hCoV-19/USA/LA-EVTL948/2020\|EPI_ISL_653639\|2020-08 | .................... | .................... | ..................... | .................... | .................... | .................... |
| S | 2940 | hCoV-19/USA/LA-EVTL949/2020\|EPI_ISL_653640\|2020-08 | .................... | .................... | ..................... | .................... | .................... | .................... |
| S | 2941 | hCoV-19/USA/LA-EVTL950/2020\|EPI_ISL_653641\|2020-08 | .................... | .................... | ..................... | .................... | .................... | .................... |
| S | 2942 | hCoV-19/USA/LA-EVTL951/2020\|EPI_ISL_653642\|2020-08 | .................... | .................... | ..................... | .................... | .................... | .................... |
| S | 2943 | hCoV-19/USA/LA-EVTL952/2020\|EPI_ISL_653643\|2020-08 | .................... | .................... | ..................... | .................... | .................... | .................... |
| S | 2944 | hCoV-19/USA/LA-EVTL953/2020\|EPI_ISL_653644\|2020-08 | .................... | .................... | ..................... | .................... | .................... | .................... |
| S | 2945 | hCoV-19/USA/LA-EVTL954/2020\|EPI_ISL_653645\|2020-08 | .................... | .................... | ..................... | .................... | .................... | .................... |
| S | 2946 | hCoV-19/USA/LA-EVTL955/2020\|EPI_ISL_653646\|2020-08 | .................... | .................... | ..................... | .................... | .................... | .................... |
| S | 2947 | hCoV-19/USA/LA-EVTL956/2020\|EPI_ISL_653647\|2020-08 | .................... | .................... | ..................... | .................... | .................... | .................... |
| S | 2948 | hCoV-19/USA/LA-EVTL957/2020\|EPI_ISL_653648\|2020-08 | .................... | .................... | ..................... | .................... | .................... | .................... |
| S | 2949 | hCoV-19/USA/LA-EVTL958/2020\|EPI_ISL_653649\|2020-08 | .................... | .................... | ..................... | .................... | .................... | .................... |
| S | 2950 | hCoV-19/USA/LA-EVTL959/2020\|EPI_ISL_653650\|2020-08 | .................... | .................... | ..................... | .................... | .................... | .................... |
| S | 2951 | hCoV-19/USA/LA-EVTL960/2020\|EPI_ISL_653651\|2020-08 | .................... | .................... | ..................... | .................... | .................... | .................... |
| S | 2952 | hCoV-19/USA/LA-EVTL961/2020\|EPI_ISL_653652\|2020-08 | .................... | .................... | ..................... | .................... | .................... | .................... |
| S | 2953 | hCoV-19/USA/LA-EVTL962/2020\|EPI_ISL_653653\|2020-08 | .................... | .................... | ..................... | .................... | .................... | .................... |
| S | 2954 | hCoV-19/USA/LA-EVTL963/2020\|EPI_ISL_653654\|2020-08 | .................... | .................... | ..................... | .................... | .................... | .................... |
| S | 2955 | hCoV-19/USA/LA-EVTL964/2020\|EPI_ISL_653655\|2020-08 | .................... | .................... | ..................... | .................... | .................... | .................... |
| S | 2956 | hCoV-19/USA/LA-EVTL965/2020\|EPI_ISL_653656\|2020-08 | .................... | .................... | ..................... | .................... | .................... | .................... |
| S | 2957 | hCoV-19/USA/LA-EVTL966/2020\|EPI_ISL_653657\|2020-08 | .................... | .................... | ..................... | .................... | .................... | .................... |
| S | 2958 | hCoV-19/USA/LA-EVTL967/2020\|EPI_ISL_653658\|2020-08 | .................... | .................... | ..................... | .................... | .................... | .................... |
| S | 2959 | hCoV-19/USA/LA-EVTL968/2020\|EPI_ISL_653659\|2020-08 | .................... | .A.................. | ..................... | .................... | .................... | .................... |
| S | 2960 | hCoV-19/USA/LA-EVTL969/2020\|EPI_ISL_653660\|2020-08 | .................... | .A.................. | ..................... | .................... | .................... | .................... |
| S | 2961 | hCoV-19/USA/LA-EVTL970/2020\|EPI_ISL_653661\|2020-08 | .................... | .................... | ..................... | .................... | .................... | .................... |
| S | 2962 | hCoV-19/USA/LA-EVTL971/2020\|EPI_ISL_653662\|2020-08 | .................... | .A.................. | ..................... | .................... | .................... | .................... |
| S | 2963 | hCoV-19/USA/LA-EVTL972/2020\|EPI_ISL_653663\|2020-08 | .................... | .................... | ..................... | .................... | .................... | .................... |
| S | 2964 | hCoV-19/USA/LA-EVTL973/2020\|EPI_ISL_653664\|2020-08 | .................... | .................... | ..................... | .................... | .................... | .................... |
| S | 2965 | hCoV-19/USA/LA-EVTL974/2020\|EPI_ISL_653665\|2020-08 | .................... | .................... | ..................... | .................... | .................... | .................... |
| S | 2966 | hCoV-19/USA/TX-DSHS-1329/2020\|EPI_ISL_654911\|2020- | .................... | .................... | ..................... | .................... | .................... | .................... |
| S | 2967 | hCoV-19/USA/TX-DSHS-1331/2020\|EPI_ISL_654913\|2020- | .................... | .................... | ..................... | .................... | .................... | .................... |
| S | 2968 | hCoV-19/USA/TX-DSHS-1345/2020\|EPI_ISL_654926\|2020- | .................... | .................... | ..................... | .................... | .................... | .................... |
| S | 2969 | hCoV-19/USA/TX-DSHS-1359/2020\|EPI_ISL_654937\|2020- | .................... | .................... | ..................... | .................... | .................... | .................... |
| S | 2970 | hCoV-19/Sweden/20-08969/2020\|EPI_ISL_654942\|2020-0 | .................... | .................... | ..................... | .................... | .................... | .................... |
| S | 2971 | hCoV-19/Sweden/20-22124/2020\|EPI_ISL_654943\|2020-0 | .................... | .................... | ..................... | .................... | .................... | .................... |
| S | 2972 | hCoV-19/Sweden/20-52720/2020\|EPI_ISL_654944\|2020-0 | .................... | .................... | ..................... | .................... | .................... | .................... |
| S | 2973 | hCoV-19/Sweden/20-52766/2020\|EPI_ISL_654945\|2020-0 | .................... | .................... | ..................... | .................... | .................... | .................... |

| | | | | | | | |
|---|---|---|---|---|---|---|---|
| S | 2974 hCoV-19/Sweden/20-52767/2020\|EPI_ISL_654946\|2020-0 | ···················· | ···················· | ····················· | ···················· | ···················· | ···················· |
| S | 2975 hCoV-19/Sweden/20-52769/2020\|EPI_ISL_654947\|2020-0 | ···················· | ···················· | ····················· | ···················· | ···················· | ···················· |
| S | 2976 hCoV-19/Sweden/20-52770/2020\|EPI_ISL_654948\|2020-0 | ···················· | ···················· | ····················· | ···················· | ···················· | ···················· |
| S | 2977 hCoV-19/Sweden/20-52774/2020\|EPI_ISL_654949\|2020-0 | ···················· | ···················· | ····················· | ···················· | ···················· | ···················· |
| S | 2978 hCoV-19/Sweden/20-52775/2020\|EPI_ISL_654950\|2020-0 | ···················· | ···················· | ····················· | ···················· | ···················· | ···················· |
| S | 2979 hCoV-19/Sweden/20-52776/2020\|EPI_ISL_654951\|2020-0 | ···················· | ···················· | ····················· | ···················· | ···················· | ···················· |
| S | 2980 hCoV-19/Sweden/20-08963/2020\|EPI_ISL_654952\|2020-0 | ···················· | ···················· | ····················· | ···················· | ···················· | ···················· |
| S | 2981 hCoV-19/Sweden/20-23086/2020\|EPI_ISL_654953\|2020-0 | ···················· | ···················· | ····················· | ···················· | ···················· | ···················· |
| S | 2982 hCoV-19/Sweden/20-52782/2020\|EPI_ISL_654954\|2020-0 | ···················· | ···················· | ····················· | ···················· | ···················· | ···················· |
| S | 2983 hCoV-19/Sweden/20-52831/2020\|EPI_ISL_654956\|2020-0 | ···················· | ···················· | ····················· | ···················· | ···················· | ···················· |
| S | 2984 hCoV-19/England/QEUH-B6C869/2020\|EPI_ISL_654959\|20 | ···················· | ···················· | ····················· | ···················· | ···················· | ···················· |
| S | 2985 hCoV-19/England/QEUH-B695CF/2020\|EPI_ISL_654960\|20 | ···················· | ···················· | ····················· | ···················· | ···················· | ···················· |
| S | 2986 hCoV-19/England/QEUH-B69F0D/2020\|EPI_ISL_654961\|20 | ···················· | ···················· | ····················· | ···················· | ···················· | ···················· |
| S | 2987 hCoV-19/England/QEUH-B685B1/2020\|EPI_ISL_654962\|20 | ···················· | ···················· | ····················· | ···················· | ···················· | ···················· |
| S | 2988 hCoV-19/England/QEUH-B681DB/2020\|EPI_ISL_654963\|20 | ···················· | ···················· | ····················· | ···················· | ···················· | ···················· |
| S | 2989 hCoV-19/England/QEUH-B68A76/2020\|EPI_ISL_654964\|20 | ···················· | ···················· | ····················· | ···················· | ···················· | ···················· |
| S | 2990 hCoV-19/England/QEUH-B6C034/2020\|EPI_ISL_654965\|20 | ···················· | ···················· | ····················· | ···················· | ···················· | ···················· |
| S | 2991 hCoV-19/England/QEUH-B6ACE7/2020\|EPI_ISL_654966\|20 | ···················· | ···················· | ····················· | ···················· | ···················· | ···················· |
| S | 2992 hCoV-19/England/QEUH-B6679F/2020\|EPI_ISL_654967\|20 | ···················· | ···················· | ····················· | ···················· | ···················· | ···················· |
| S | 2993 hCoV-19/England/QEUH-B669B7/2020\|EPI_ISL_654968\|20 | ···················· | ···················· | ····················· | ···················· | ···················· | ···················· |
| S | 2994 hCoV-19/England/QEUH-B6A33D/2020\|EPI_ISL_654969\|20 | ···················· | ···················· | ····················· | ···················· | ···················· | ···················· |
| S | 2995 hCoV-19/England/QEUH-B68B37/2020\|EPI_ISL_654970\|20 | ···················· | ···················· | ····················· | ···················· | ···················· | ···················· |
| S | 2996 hCoV-19/England/QEUH-B69EB5/2020\|EPI_ISL_654971\|20 | ···················· | ···················· | ····················· | ···················· | ···················· | ···················· |
| S | 2997 hCoV-19/England/QEUH-B687CA/2020\|EPI_ISL_654972\|20 | ···················· | ···················· | ····················· | ···················· | ···················· | ···················· |
| S | 2998 hCoV-19/England/QEUH-B69608/2020\|EPI_ISL_654973\|20 | ···················· | ···················· | ····················· | ···················· | ···················· | ···················· |
| S | 2999 hCoV-19/England/QEUH-B65CFB/2020\|EPI_ISL_654974\|20 | ···················· | ···················· | ····················· | ···················· | ···················· | ···················· |
| S | 3000 hCoV-19/England/QEUH-B66753/2020\|EPI_ISL_654975\|20 | ···················· | ···················· | ····················· | ···················· | ···················· | ···················· |
| S | 3001 hCoV-19/England/QEUH-B69F58/2020\|EPI_ISL_654976\|20 | ···················· | ···················· | ····················· | ···················· | ···················· | ···················· |
| S | 3002 hCoV-19/England/QEUH-B696BD/2020\|EPI_ISL_654977\|20 | ···················· | ···················· | ····················· | ···················· | ···················· | ···················· |
| S | 3003 hCoV-19/England/QEUH-B6BD7A/2020\|EPI_ISL_654978\|20 | ···················· | ···················· | ····················· | ···················· | ···················· | ···················· |
| S | 3004 hCoV-19/England/QEUH-B684D2/2020\|EPI_ISL_654979\|20 | ···················· | ···················· | ····················· | ···················· | ···················· | ···················· |
| S | 3005 hCoV-19/England/QEUH-B6AD11/2020\|EPI_ISL_654980\|20 | ···················· | ···················· | ····················· | ···················· | ···················· | ···················· |
| S | 3006 hCoV-19/England/QEUH-B66A2D/2020\|EPI_ISL_654981\|20 | ···················· | ·········A·········· | ····················· | ···················· | ······T············· | ···················· |
| S | 3007 hCoV-19/England/QEUH-B69653/2020\|EPI_ISL_654982\|20 | ···················· | ···················· | ····················· | ···················· | ···················· | ···················· |
| S | 3008 hCoV-19/England/QEUH-B66A69/2020\|EPI_ISL_654983\|20 | ···················· | ···················· | ····················· | ···················· | ···················· | ···················· |
| S | 3009 hCoV-19/England/QEUH-B65CCE/2020\|EPI_ISL_654984\|20 | ···················· | ···················· | ····················· | ···················· | ···················· | ···················· |
| S | 3010 hCoV-19/England/QEUH-B69714/2020\|EPI_ISL_654985\|20 | ···················· | ···················· | ····················· | ···················· | ···················· | ···················· |
| S | 3011 hCoV-19/England/QEUH-B69538/2020\|EPI_ISL_654986\|20 | ···················· | ·A·················· | ····················· | ···················· | ···················· | ···A················ |
| S | 3012 hCoV-19/England/QEUH-B6ACF6/2020\|EPI_ISL_654987\|20 | ···················· | ···················· | ····················· | ···················· | ···················· | ···················· |
| S | 3013 hCoV-19/England/QEUH-B6698A/2020\|EPI_ISL_654988\|20 | ···················· | ···················· | ····················· | ···················· | ······T············· | ···················· |
| S | 3014 hCoV-19/England/QEUH-B65D43/2020\|EPI_ISL_654989\|20 | ···················· | ···················· | ····················· | ···················· | ···················· | ···················· |
| S | 3015 hCoV-19/England/QEUH-B6C8A5/2020\|EPI_ISL_654990\|20 | ···················· | ···················· | ····················· | ···················· | ···················· | ···················· |
| S | 3016 hCoV-19/England/QEUH-B66A87/2020\|EPI_ISL_654991\|20 | ···················· | ···················· | ····················· | ···················· | ···················· | ···················· |
| S | 3017 hCoV-19/England/QEUH-B69E4C/2020\|EPI_ISL_654992\|20 | ···················· | ···················· | ····················· | ···················· | ···················· | ···················· |
| S | 3018 hCoV-19/England/QEUH-B669D5/2020\|EPI_ISL_654993\|20 | ···················· | ···················· | ····················· | ···················· | ···················· | ···················· |
| S | 3019 hCoV-19/England/QEUH-B68AD0/2020\|EPI_ISL_654994\|20 | ···················· | ···················· | ····················· | ···················· | ···················· | ···················· |
| S | 3020 hCoV-19/England/QEUH-B6C386/2020\|EPI_ISL_654995\|20 | ···················· | ···················· | ····················· | ···················· | ···················· | ···················· |
| S | 3021 hCoV-19/England/QEUH-B669E4/2020\|EPI_ISL_654996\|20 | ···················· | ···················· | ····················· | ···················· | ······T············· | ···················· |
| S | 3022 hCoV-19/England/QEUH-B437E3/2020\|EPI_ISL_657109\|20 | ···················· | ···················· | ····················· | ···················· | ···················· | ···················· |
| S | 3023 hCoV-19/England/QEUH-B43464/2020\|EPI_ISL_657114\|20 | ···················· | ···················· | ····················· | ···················· | ···················· | ···················· |
| S | 3024 hCoV-19/England/QEUH-B42553/2020\|EPI_ISL_657120\|20 | ···················· | ···················· | ····················· | ···················· | ···················· | ···················· |
| S | 3025 hCoV-19/England/QEUH-B424A1/2020\|EPI_ISL_657121\|20 | ···················· | ···················· | ····················· | ···················· | ···················· | ···················· |
| S | 3026 hCoV-19/England/QEUH-B402A9/2020\|EPI_ISL_657122\|20 | ···················· | ···················· | ····················· | ···················· | ···················· | ···················· |
| S | 3027 hCoV-19/England/QEUH-B42368/2020\|EPI_ISL_657123\|20 | ···················· | ···················· | ····················· | ···················· | ···················· | ···················· |
| S | 3028 hCoV-19/England/QEUH-B40591/2020\|EPI_ISL_657124\|20 | ···················· | ···················· | ····················· | ···················· | ···················· | ···················· |
| S | 3029 hCoV-19/England/QEUH-B434DD/2020\|EPI_ISL_657125\|20 | ···················· | ···················· | ····················· | ···················· | ···················· | ···················· |
| S | 3030 hCoV-19/England/QEUH-B43640/2020\|EPI_ISL_657126\|20 | ···················· | ···················· | ····················· | ···················· | ···················· | ···················· |
| S | 3031 hCoV-19/England/QEUH-B4216E/2020\|EPI_ISL_657127\|20 | ···················· | ·······K············ | ····················· | ···················· | ···················· | ···················· |
| S | 3032 hCoV-19/England/QEUH-B401F7/2020\|EPI_ISL_657128\|20 | ···················· | ···················· | ····················· | ···················· | ···················· | ···················· |
| S | 3033 hCoV-19/England/QEUH-B40995/2020\|EPI_ISL_657129\|20 | ···················· | ···················· | ····················· | ···················· | ···················· | ···················· |
| S | 3034 hCoV-19/England/QEUH-B40740/2020\|EPI_ISL_657190\|20 | ···················· | ···················· | ····················· | ···················· | ···················· | ···················· |
| S | 3035 hCoV-19/England/QEUH-B43956/2020\|EPI_ISL_657192\|20 | ···················· | ···················· | ····················· | ···················· | ···················· | ···················· |
| S | 3036 hCoV-19/England/QEUH-B42447/2020\|EPI_ISL_657193\|20 | ···················· | ···················· | ····················· | ···················· | ···················· | ···················· |
| S | 3037 hCoV-19/England/QEUH-B4232C/2020\|EPI_ISL_657194\|20 | ········T··········· | ···················· | ····················· | ···················· | ···················· | ···················· |
| S | 3038 hCoV-19/England/QEUH-B40713/2020\|EPI_ISL_657195\|20 | ···················· | ···················· | ····················· | ···················· | ···················· | ···················· |
| S | 3039 hCoV-19/England/QEUH-B435BC/2020\|EPI_ISL_657196\|20 | ···················· | ···················· | ····················· | ···················· | ···················· | ···················· |
| S | 3040 hCoV-19/England/QEUH-B3FE5E/2020\|EPI_ISL_657197\|20 | ···················· | ···················· | ····················· | ···················· | ···················· | ···················· |
| S | 3041 hCoV-19/England/QEUH-B40072/2020\|EPI_ISL_657198\|20 | ···················· | ···················· | ····················· | ···················· | ···················· | ···················· |
| S | 3042 hCoV-19/England/QEUH-B421D7/2020\|EPI_ISL_657199\|20 | ···················· | ···················· | ····················· | ···················· | ···················· | ···················· |
| S | 3043 hCoV-19/England/QEUH-B3F708/2020\|EPI_ISL_658445\|20 | ···················· | ···················· | ····················· | ···················· | ···················· | ···················· |
| S | 3044 hCoV-19/England/QEUH-B3FB57/2020\|EPI_ISL_658446\|20 | ···················· | ···················· | ····················· | ···················· | ···················· | ···················· |
| S | 3045 hCoV-19/England/QEUH-B40FC0/2020\|EPI_ISL_658450\|20 | ···················· | ···················· | ····················· | ···················· | ···················· | ···················· |
| S | 3046 hCoV-19/England/QEUH-B3FC09/2020\|EPI_ISL_658451\|20 | ···················· | ···················· | ····················· | ···················· | ···················· | ···················· |
| S | 3047 hCoV-19/England/QEUH-B3FA96/2020\|EPI_ISL_658452\|20 | ···················· | ···················· | ····················· | ···················· | ···················· | ···················· |
| S | 3048 hCoV-19/England/QEUH-B3FC90/2020\|EPI_ISL_658453\|20 | ···················· | ···················· | ····················· | ···················· | ···················· | ···················· |
| S | 3049 hCoV-19/England/QEUH-B3F216/2020\|EPI_ISL_658456\|20 | ···················· | ···················· | ····················· | ···················· | ···················· | ···················· |
| S | 3050 hCoV-19/England/QEUH-B3F8BA/2020\|EPI_ISL_658457\|20 | ···················· | ···················· | ····················· | ···················· | ···················· | ···················· |
| S | 3051 hCoV-19/England/QEUH-B3F832/2020\|EPI_ISL_658458\|20 | ···················· | ···················· | ····················· | ···················· | ···················· | ···················· |
| S | 3052 hCoV-19/England/QEUH-B412F3/2020\|EPI_ISL_658459\|20 | ···················· | ···················· | ····················· | ···················· | ···················· | ···················· |
| S | 3053 hCoV-19/England/QEUH-B302E8/2020\|EPI_ISL_659776\|20 | ···················· | ···················· | ····················· | ···················· | ···················· | ···················· |

| | | | | | | | |
|---|---|---|---|---|---|---|---|
| S | 3054 hCoV-19/England/QEUH-B2CD75/2020\|EPI_ISL_659781\|20 | ···················· | ···················· | ···················· | ···················· | ···················· | ···················· |
| S | 3055 hCoV-19/England/QEUH-B2EF31/2020\|EPI_ISL_659782\|20 | ···················· | ···················· | ···················· | ···················· | ···················· | ···················· |
| S | 3056 hCoV-19/England/QEUH-B2F5E3/2020\|EPI_ISL_659783\|20 | ···················· | ···················· | ···················· | ···················· | ···················· | ···················· |
| S | 3057 hCoV-19/England/QEUH-B2F668/2020\|EPI_ISL_659784\|20 | ···················· | ···················· | ···················· | ···················· | ···················· | ···················· |
| S | 3058 hCoV-19/England/QEUH-B2D521/2020\|EPI_ISL_659787\|20 | ···················· | ···················· | ···················· | ···················· | ···················· | ···················· |
| S | 3059 hCoV-19/England/QEUH-B2DF9C/2020\|EPI_ISL_659789\|20 | ···················· | ···················· | ···················· | ···················· | ···················· | ···················· |
| S | 3060 hCoV-19/England/204380511/2020\|EPI_ISL_660750\|2020 | ···················· | ···················· | ···················· | ···················· | ···················· | ···················· |
| S | 3061 hCoV-19/England/204440445/2020\|EPI_ISL_660752\|2020 | ···················· | ···················· | ···················· | ···················· | ···················· | ···················· |
| S | 3062 hCoV-19/England/204480236/2020\|EPI_ISL_660753\|2020 | ···················· | ···················· | ···················· | ···················· | ···················· | ···················· |
| S | 3063 hCoV-19/England/204490922/2020\|EPI_ISL_660758\|2020 | ···················· | ···················· | ···················· | ···················· | ···················· | ···················· |
| S | 3064 hCoV-19/England/204490923/2020\|EPI_ISL_660759\|2020 | ···················· | ···················· | ···················· | ···················· | ···················· | ···················· |
| S | 3065 hCoV-19/England/204490931/2020\|EPI_ISL_660761\|2020 | ···················· | ···················· | ···················· | ···················· | ···················· | ···················· |
| S | 3066 hCoV-19/England/204490933/2020\|EPI_ISL_660762\|2020 | ···················· | ···················· | ···················· | ···················· | ···················· | ···················· |
| S | 3067 hCoV-19/England/204490935/2020\|EPI_ISL_660763\|2020 | ···················· | ···················· | ···················· | ···················· | ···················· | ···················· |
| S | 3068 hCoV-19/England/204490945/2020\|EPI_ISL_660764\|2020 | ···················· | ·A·················· | ···················· | ···················· | ···················· | ···················· |
| S | 3069 hCoV-19/England/204490969/2020\|EPI_ISL_660765\|2020 | ···················· | ···················· | ···················· | ···················· | ···················· | ···················· |
| S | 3070 hCoV-19/England/204491152/2020\|EPI_ISL_660767\|2020 | ···················· | ···················· | ···················· | ···················· | ···················· | ···················· |
| S | 3071 hCoV-19/England/204491176/2020\|EPI_ISL_660770\|2020 | ···················· | ···················· | ···················· | ···················· | ···················· | ···················· |
| S | 3072 hCoV-19/England/204491198/2020\|EPI_ISL_660771\|2020 | ···················· | ···················· | ···················· | ···················· | ···················· | ···················· |
| S | 3073 hCoV-19/England/MILK-B05A02/2020\|EPI_ISL_652214\|20 | ···················· | ···················· | ···················· | ···················· | ···················· | ···················· |
| S | 3074 hCoV-19/England/MILK-B05A3F/2020\|EPI_ISL_652215\|20 | ···················· | ···················· | ···················· | ···················· | ···················· | ···················· |
| S | 3075 hCoV-19/England/MILK-B05A99/2020\|EPI_ISL_652216\|20 | ···················· | ·A·······A·········· | ···················· | ···················· | ···················· | ···················· |
| S | 3076 hCoV-19/England/MILK-B05AA8/2020\|EPI_ISL_652217\|20 | ···················· | ···················· | ···················· | ···················· | ···················· | ···················· |
| S | 3077 hCoV-19/England/MILK-B05BC3/2020\|EPI_ISL_652218\|20 | ···················· | ···················· | ···················· | ···················· | ···················· | ···················· |
| S | 3078 hCoV-19/England/MILK-B05BF0/2020\|EPI_ISL_652219\|20 | ···················· | ···················· | ···················· | ···················· | ···················· | ···················· |
| S | 3079 hCoV-19/England/MILK-B05C1B/2020\|EPI_ISL_652220\|20 | ···················· | ···················· | ···················· | ···················· | ···················· | ···················· |
| S | 3080 hCoV-19/England/MILK-B05C48/2020\|EPI_ISL_652221\|20 | ···················· | ···················· | ···················· | ···················· | ······T············· | ···················· |
| S | 3081 hCoV-19/England/MILK-B05C57/2020\|EPI_ISL_652222\|20 | ···················· | ···················· | ···················· | ···················· | ···················· | ···················· |
| S | 3082 hCoV-19/England/MILK-B05CEE/2020\|EPI_ISL_652223\|20 | ···················· | ···················· | ···················· | ···················· | ···················· | ···················· |
| S | 3083 hCoV-19/England/MILK-B05D09/2020\|EPI_ISL_652224\|20 | ···················· | ···················· | ···················· | ···················· | ···················· | ···················· |
| S | 3084 hCoV-19/England/MILK-B05D36/2020\|EPI_ISL_652225\|20 | ···················· | ···················· | ···················· | ···················· | ···················· | ···················· |
| S | 3085 hCoV-19/England/MILK-B05D45/2020\|EPI_ISL_652226\|20 | ···················· | ···················· | ···················· | ···················· | ···················· | ···················· |
| S | 3086 hCoV-19/England/MILK-B063BB/2020\|EPI_ISL_652227\|20 | ···················· | ···················· | ···················· | ···················· | ···················· | ···················· |
| S | 3087 hCoV-19/Wales/MILK-B063CA/2020\|EPI_ISL_652228\|2020 | ···················· | ···················· | ···················· | ···················· | ···················· | ···················· |
| S | 3088 hCoV-19/Wales/MILK-B063E8/2020\|EPI_ISL_652229\|2020 | ···················· | ···················· | ···················· | ···················· | ···················· | ···················· |
| S | 3089 hCoV-19/Wales/MILK-B063F7/2020\|EPI_ISL_652230\|2020 | ···················· | ···················· | ···················· | ···················· | ···················· | ···················· |
| S | 3090 hCoV-19/England/MILK-B06403/2020\|EPI_ISL_652231\|20 | ···················· | ···················· | ···················· | ···················· | ···················· | ···················· |
| S | 3091 hCoV-19/England/MILK-B0644F/2020\|EPI_ISL_652232\|20 | ···················· | ···················· | ···················· | ···················· | ···················· | ···················· |
| S | 3092 hCoV-19/Wales/MILK-B0646D/2020\|EPI_ISL_652233\|2020 | ···················· | ···················· | ···················· | ···················· | ···················· | ···················· |
| S | 3093 hCoV-19/Wales/MILK-B0647C/2020\|EPI_ISL_652234\|2020 | ···················· | ···················· | ···················· | ···················· | ···················· | ···················· |
| S | 3094 hCoV-19/Wales/MILK-B064E5/2020\|EPI_ISL_652235\|2020 | ···················· | ···················· | ···················· | ···················· | ···················· | ···················· |
| S | 3095 hCoV-19/Wales/MILK-B064F4/2020\|EPI_ISL_652236\|2020 | ···················· | ···················· | ···················· | ···················· | ···················· | ···················· |
| S | 3096 hCoV-19/England/MILK-B0652E/2020\|EPI_ISL_652237\|20 | ···················· | ···················· | ···················· | ···················· | ···················· | ···················· |
| S | 3097 hCoV-19/England/MILK-B0653D/2020\|EPI_ISL_652238\|20 | ···················· | ···················· | ···················· | ···················· | ···················· | ···················· |
| S | 3098 hCoV-19/England/MILK-B06597/2020\|EPI_ISL_652239\|20 | ···················· | ···················· | ···················· | ···················· | ···················· | ···················· |
| S | 3099 hCoV-19/England/MILK-B065E2/2020\|EPI_ISL_652240\|20 | ···················· | ···················· | ···················· | ···················· | ···················· | ···················· |
| S | 3100 hCoV-19/England/MILK-B06649/2020\|EPI_ISL_652241\|20 | ···················· | ···················· | ···················· | ···················· | ···················· | ···················· |
| S | 3101 hCoV-19/England/MILK-B06667/2020\|EPI_ISL_652242\|20 | ···················· | ···················· | ···················· | ···················· | ···················· | ···················· |
| S | 3102 hCoV-19/England/MILK-B066A3/2020\|EPI_ISL_652243\|20 | ···················· | ···················· | ···················· | ···················· | ···················· | ···················· |
| S | 3103 hCoV-19/England/MILK-B0670A/2020\|EPI_ISL_652244\|20 | ···················· | ···················· | ···················· | ···················· | ···················· | ···················· |
| S | 3104 hCoV-19/England/MILK-B06764/2020\|EPI_ISL_652245\|20 | ···················· | ···················· | ···················· | ···················· | ···················· | ···················· |
| S | 3105 hCoV-19/England/MILK-B06773/2020\|EPI_ISL_652246\|20 | ···················· | ···················· | ···················· | ···················· | ···················· | ···················· |
| S | 3106 hCoV-19/England/MILK-B067BF/2020\|EPI_ISL_652247\|20 | ···················· | ···················· | ···················· | ···················· | ···················· | ···················· |
| S | 3107 hCoV-19/England/MILK-B067CE/2020\|EPI_ISL_652248\|20 | ···················· | ···················· | ···················· | ···················· | ···················· | ···················· |
| S | 3108 hCoV-19/England/MILK-B06807/2020\|EPI_ISL_652249\|20 | ···················· | ···················· | ···················· | ···················· | ···················· | ···················· |
| S | 3109 hCoV-19/England/MILK-B06843/2020\|EPI_ISL_652250\|20 | ···················· | ···················· | ···················· | ···················· | ···················· | ···················· |
| S | 3110 hCoV-19/England/MILK-B06870/2020\|EPI_ISL_652251\|20 | ···················· | ···················· | ···················· | ···················· | ···················· | ···················· |
| S | 3111 hCoV-19/England/MILK-B068CB/2020\|EPI_ISL_652252\|20 | ···················· | ···················· | ···················· | ···················· | ···················· | ···················· |
| S | 3112 hCoV-19/England/MILK-B06922/2020\|EPI_ISL_652253\|20 | ···················· | ···················· | ···················· | ···················· | ······T············· | ···················· |
| S | 3113 hCoV-19/England/MILK-B0695F/2020\|EPI_ISL_652254\|20 | ···················· | ···················· | ···················· | ···················· | ······T············· | ···················· |
| S | 3114 hCoV-19/England/MILK-B0696E/2020\|EPI_ISL_652255\|20 | ···················· | ···················· | ···················· | ···················· | ···················· | ···················· |
| S | 3115 hCoV-19/England/MILK-B0697D/2020\|EPI_ISL_652256\|20 | ···················· | ···················· | ···················· | ···················· | ···················· | ···················· |
| S | 3116 hCoV-19/England/MILK-B0698C/2020\|EPI_ISL_652257\|20 | ···················· | ···················· | ···················· | ···················· | ···················· | ···················· |
| S | 3117 hCoV-19/England/MILK-B06C65/2020\|EPI_ISL_652269\|20 | ···················· | ···················· | ···················· | ···················· | ···················· | ···················· |
| S | 3118 hCoV-19/England/MILK-B06D9F/2020\|EPI_ISL_652276\|20 | ···················· | ···················· | ···················· | ···················· | ···················· | ···················· |
| S | 3119 hCoV-19/England/MILK-B06DBD/2020\|EPI_ISL_652277\|20 | ···················· | ···················· | ···················· | ···················· | ···················· | ···················· |
| S | 3120 hCoV-19/England/MILK-B06E14/2020\|EPI_ISL_652278\|20 | ···················· | ···················· | ···················· | ···················· | ···················· | ···················· |
| S | 3121 hCoV-19/England/MILK-B06E50/2020\|EPI_ISL_652279\|20 | ···················· | ···················· | ···················· | ···················· | ···················· | ···················· |
| S | 3122 hCoV-19/USA/LA-EVTL854/2020\|EPI_ISL_653545\|2020-07 | ···················· | ···················· | ···················· | ···················· | ···················· | ···················· |
| S | 3123 hCoV-19/USA/LA-EVTL855/2020\|EPI_ISL_653546\|2020-06 | ···················· | ···················· | ···················· | ···················· | ···················· | ···················· |
| S | 3124 hCoV-19/USA/LA-EVTL856/2020\|EPI_ISL_653547\|2020-06 | ···················· | ···················· | ···················· | ···················· | ···················· | ···················· |
| S | 3125 hCoV-19/USA/LA-EVTL857/2020\|EPI_ISL_653548\|2020-07 | ···················· | ···················· | ···················· | ···················· | ···················· | ···················· |
| S | 3126 hCoV-19/USA/LA-EVTL858/2020\|EPI_ISL_653549\|2020-07 | ···················· | ···················· | ···················· | ···················· | ···················· | ···················· |
| S | 3127 hCoV-19/USA/LA-EVTL859/2020\|EPI_ISL_653550\|2020-07 | ···················· | ···················· | ···················· | ···················· | ···················· | ···················· |
| S | 3128 hCoV-19/USA/LA-EVTL860/2020\|EPI_ISL_653551\|2020-06 | ···················· | ···················· | ···················· | ···················· | ···················· | ···················· |
| S | 3129 hCoV-19/USA/LA-EVTL861/2020\|EPI_ISL_653552\|2020-07 | ···················· | ···················· | ···················· | ···················· | ···················· | ···················· |
| S | 3130 hCoV-19/USA/LA-EVTL862/2020\|EPI_ISL_653553\|2020-07 | ···················· | ···················· | ···················· | ···················· | ·····C·············· | ···················· |
| S | 3131 hCoV-19/USA/LA-EVTL863/2020\|EPI_ISL_653554\|2020-06 | ···················· | ·A·················· | ···················· | ···················· | ···················· | ···················· |
| S | 3132 hCoV-19/USA/LA-EVTL865/2020\|EPI_ISL_653556\|2020-07 | ···················· | ···················· | ···················· | ···················· | ···················· | ···················· |
| S | 3133 hCoV-19/USA/LA-EVTL866/2020\|EPI_ISL_653557\|2020-07 | ···················· | ···················· | ···················· | ···················· | ···················· | ···················· |

| | | | | | | | |
|---|---|---|---|---|---|---|---|
| S | 3134 hCoV-19/USA/LA-EVTL867/2020\|EPI_ISL_653558\|2020-07 | ···················· | ···················· | ···················· | ···················· | ···················· | ···················· |
| S | 3135 hCoV-19/USA/LA-EVTL868/2020\|EPI_ISL_653559\|2020-07 | ···················· | ···················· | ···················· | ···················· | ···················· | ···················· |
| S | 3136 hCoV-19/USA/LA-EVTL869/2020\|EPI_ISL_653560\|2020-07 | ···················· | ···················· | ···················· | ···················· | ···················· | ···················· |
| S | 3137 hCoV-19/USA/LA-EVTL870/2020\|EPI_ISL_653561\|2020-07 | ···················· | ···················· | ···················· | ···················· | ···················· | ···················· |
| S | 3138 hCoV-19/USA/LA-EVTL871/2020\|EPI_ISL_653562\|2020-07 | ···················· | ···················· | ···················· | ···················· | ···················· | ···················· |
| S | 3139 hCoV-19/USA/LA-EVTL872/2020\|EPI_ISL_653563\|2020-07 | ···················· | ···················· | ···················· | ···················· | ···················· | ···················· |
| S | 3140 hCoV-19/USA/LA-EVTL873/2020\|EPI_ISL_653564\|2020-07 | ···················· | ···················· | ···················· | ···················· | ···················· | ···················· |
| S | 3141 hCoV-19/USA/LA-EVTL874/2020\|EPI_ISL_653565\|2020-07 | ···················· | ···················· | ···················· | ···················· | ···················· | ···················· |
| S | 3142 hCoV-19/USA/LA-EVTL875/2020\|EPI_ISL_653566\|2020-07 | ···················· | ···················· | ···················· | ···················· | ···················· | ···················· |
| S | 3143 hCoV-19/USA/LA-EVTL876/2020\|EPI_ISL_653567\|2020-07 | ···················· | ···················· | ···················· | ···················· | ···················· | ···················· |
| S | 3144 hCoV-19/USA/LA-EVTL877/2020\|EPI_ISL_653568\|2020-07 | ···················· | ···················· | ···················· | ···················· | ···················· | ···················· |
| S | 3145 hCoV-19/USA/LA-EVTL878/2020\|EPI_ISL_653569\|2020-07 | ···················· | ···················· | ···················· | ···················· | ···················· | ···················· |
| S | 3146 hCoV-19/USA/LA-EVTL879/2020\|EPI_ISL_653570\|2020-07 | ···················· | ···················· | ···················· | ···················· | ···················· | ···················· |
| S | 3147 hCoV-19/USA/LA-EVTL880/2020\|EPI_ISL_653571\|2020-07 | ···················· | ···················· | ···················· | ···················· | ···················· | ···················· |
| S | 3148 hCoV-19/USA/LA-EVTL881/2020\|EPI_ISL_653572\|2020-07 | ···················· | ···················· | ···················· | ···················· | ···················· | ···················· |
| S | 3149 hCoV-19/USA/LA-EVTL882/2020\|EPI_ISL_653573\|2020-07 | ···················· | ···················· | ···················· | ···················· | ···················· | ···················· |
| S | 3150 hCoV-19/USA/LA-EVTL883/2020\|EPI_ISL_653574\|2020-07 | ···················· | ···················· | ···················· | ···················· | ···················· | ···················· |
| S | 3151 hCoV-19/USA/LA-EVTL884/2020\|EPI_ISL_653575\|2020-07 | ···················· | ···················· | ···················· | ···················· | ···················· | ···················· |
| S | 3152 hCoV-19/USA/LA-EVTL885/2020\|EPI_ISL_653576\|2020-07 | ···················· | ···················· | ···················· | ···················· | ···················· | ···················· |
| S | 3153 hCoV-19/USA/LA-EVTL886/2020\|EPI_ISL_653577\|2020-07 | ···················· | ···················· | ···················· | ···················· | ···················· | ···················· |
| S | 3154 hCoV-19/USA/LA-EVTL887/2020\|EPI_ISL_653578\|2020-07 | ···················· | ···················· | ···················· | ···················· | ···················· | ···················· |
| S | 3155 hCoV-19/USA/LA-EVTL888/2020\|EPI_ISL_653579\|2020-07 | ···················· | ···················· | ···················· | ···················· | ···················· | ···················· |
| S | 3156 hCoV-19/USA/LA-EVTL889/2020\|EPI_ISL_653580\|2020-07 | ···················· | ···················· | ···················· | ···················· | ···················· | ···················· |
| S | 3157 hCoV-19/USA/LA-EVTL890/2020\|EPI_ISL_653581\|2020-07 | ···················· | ···················· | ···················· | ···················· | ···················· | ···················· |
| S | 3158 hCoV-19/USA/LA-EVTL891/2020\|EPI_ISL_653582\|2020-07 | ···················· | ···················· | ···················· | ···················· | ···················· | ···················· |
| S | 3159 hCoV-19/USA/LA-EVTL892/2020\|EPI_ISL_653583\|2020-07 | ···················· | ···················· | ···················· | ···················· | ···················· | ···················· |
| S | 3160 hCoV-19/USA/LA-EVTL893/2020\|EPI_ISL_653584\|2020-07 | ···················· | ···················· | ···················· | ···················· | ···················· | ···················· |
| S | 3161 hCoV-19/USA/LA-EVTL894/2020\|EPI_ISL_653585\|2020-07 | ···················· | ···················· | ···················· | ···················· | ···················· | ···················· |
| S | 3162 hCoV-19/USA/LA-EVTL895/2020\|EPI_ISL_653586\|2020-07 | ···················· | ···················· | ···················· | ···················· | ···················· | ···················· |
| S | 3163 hCoV-19/USA/LA-EVTL896/2020\|EPI_ISL_653587\|2020-07 | ···················· | ···················· | ···················· | ···················· | ···················· | ···················· |
| S | 3164 hCoV-19/USA/LA-EVTL897/2020\|EPI_ISL_653588\|2020-07 | ···················· | ···················· | ···················· | ···················· | ···················· | ···················· |
| S | 3165 hCoV-19/Vietnam/PIHCM-408/2020\|EPI_ISL_654876\|2020 | ···················· | ···················· | ···················· | ···················· | ···················· | ···················· |
| S | 3166 hCoV-19/Vietnam/PIHCM-237/2020\|EPI_ISL_654877\|2020 | ···················· | ···················· | ···················· | ···················· | ···················· | ···················· |
| S | 3167 hCoV-19/Vietnam/PIHCM-305/2020\|EPI_ISL_654878\|2020 | ···················· | ···················· | ···················· | ···················· | ···················· | ···················· |
| S | 3168 hCoV-19/Vietnam/PIHCM-506/2020\|EPI_ISL_654879\|2020 | ···················· | ···················· | ···················· | ···················· | ···················· | ···················· |
| S | 3169 hCoV-19/Vietnam/PIHCM-638/2020\|EPI_ISL_654881\|2020 | ···················· | ···················· | ···················· | ···················· | ···················· | ···················· |
| S | 3170 hCoV-19/Vietnam/PIHCM-371/2020\|EPI_ISL_654882\|2020 | ···················· | ···················· | ···················· | ···················· | ···················· | ···················· |
| S | 3171 hCoV-19/Vietnam/PIHCM-503/2020\|EPI_ISL_654883\|2020 | ···················· | ···················· | ···················· | ···················· | ···················· | ···················· |
| S | 3172 hCoV-19/Vietnam/PIHCM-420/2020\|EPI_ISL_654884\|2020 | ···················· | ···················· | ···················· | ···················· | ···················· | ···················· |
| S | 3173 hCoV-19/Vietnam/PIHCM-276/2020\|EPI_ISL_654885\|2020 | ···················· | ···················· | ···················· | ···················· | ···················· | ···················· |
| S | 3174 hCoV-19/Vietnam/PIHCM-462/2020\|EPI_ISL_654886\|2020 | ···················· | ···················· | ···················· | ···················· | ···················· | ···················· |
| S | 3175 hCoV-19/Vietnam/PIHCM-634/2020\|EPI_ISL_654887\|2020 | ···················· | ···················· | ···················· | ···················· | ···················· | ···················· |
| S | 3176 hCoV-19/Vietnam/PIHCM-320/2020\|EPI_ISL_654888\|2020 | ···················· | ····A··············· | ···················· | ···················· | ···················· | ···················· |
| S | 3177 hCoV-19/Vietnam/PIHCM-457/2020\|EPI_ISL_654889\|2020 | ···················· | ···················· | ···················· | ···················· | ···················· | ···················· |
| S | 3178 hCoV-19/Vietnam/22/2020\|EPI_ISL_654890\|2020-03-08 | ···················· | ···················· | ···················· | ···················· | ···················· | ···················· |
| S | 3179 hCoV-19/Vietnam/36/2020\|EPI_ISL_654891\|2020-03-11 | ···················· | ···················· | ···················· | ···················· | ···················· | ···················· |
| S | 3180 hCoV-19/Sweden/20-08972/2020\|EPI_ISL_654892\|2020-0 | ···················· | ···················· | ···················· | ···················· | ···················· | ···················· |
| S | 3181 hCoV-19/Sweden/20-22278/2020\|EPI_ISL_654893\|2020-0 | ···················· | ···················· | ···················· | ···················· | ···················· | ···················· |
| S | 3182 hCoV-19/Sweden/20-22279/2020\|EPI_ISL_654894\|2020-0 | ···················· | ···················· | ···················· | ···················· | ···················· | ···················· |
| S | 3183 hCoV-19/Sweden/20-22280/2020\|EPI_ISL_654895\|2020-0 | ···················· | ···················· | ···················· | ···················· | ···················· | ···················· |
| S | 3184 hCoV-19/Sweden/20-22281/2020\|EPI_ISL_654896\|2020-0 | ···················· | ···················· | ···················· | ···················· | ···················· | ···················· |
| S | 3185 hCoV-19/Sweden/20-22282/2020\|EPI_ISL_654897\|2020-0 | ···················· | ···················· | ···················· | ···················· | ···················· | ···················· |
| S | 3186 hCoV-19/Sweden/20-22284/2020\|EPI_ISL_654898\|2020-0 | ···················· | ···················· | ···················· | ···················· | ···················· | ···················· |
| S | 3187 hCoV-19/Sweden/20-52716/2020\|EPI_ISL_654899\|2020-0 | ···················· | ···················· | ···················· | ···················· | ···················· | ···················· |
| S | 3188 hCoV-19/England/QEUH-B408C5/2020\|EPI_ISL_657053\|20 | ···················· | ·A·················· | ···················· | ···················· | ···················· | ···················· |
| S | 3189 hCoV-19/England/QEUH-B40458/2020\|EPI_ISL_657054\|20 | ···················· | ···················· | ···················· | ···················· | ···················· | ···················· |
| S | 3190 hCoV-19/England/QEUH-B43561/2020\|EPI_ISL_657055\|20 | ···················· | ···················· | ···················· | ···················· | ···················· | ···················· |
| S | 3191 hCoV-19/England/QEUH-B4083E/2020\|EPI_ISL_657056\|20 | ···················· | ···················· | ···················· | ···················· | ···················· | ···················· |
| S | 3192 hCoV-19/England/QEUH-B401D9/2020\|EPI_ISL_657100\|20 | ···················· | ···················· | ···················· | ···················· | ···················· | ···················· |
| S | 3193 hCoV-19/England/QEUH-B42201/2020\|EPI_ISL_657102\|20 | ···················· | ···················· | ···················· | ···················· | ···················· | ···················· |
| S | 3194 hCoV-19/England/QEUH-B433FE/2020\|EPI_ISL_657103\|20 | ···················· | ···················· | ···················· | ···················· | ···················· | ···················· |
| S | 3195 hCoV-19/England/QEUH-B400CD/2020\|EPI_ISL_657104\|20 | ···················· | ···················· | ···················· | ···················· | ······T············· | ···················· |
| S | 3196 hCoV-19/England/QEUH-B401E8/2020\|EPI_ISL_657105\|20 | ···················· | ···················· | ···················· | ···················· | ···················· | ···················· |
| S | 3197 hCoV-19/England/QEUH-B404C1/2020\|EPI_ISL_657106\|20 | ···················· | ···················· | ···················· | ···················· | ···················· | ···················· |
| S | 3198 hCoV-19/England/QEUH-B432E2/2020\|EPI_ISL_657107\|20 | ···················· | ···················· | ···················· | ···················· | ···················· | ···················· |
| S | 3199 hCoV-19/England/QEUH-B40801/2020\|EPI_ISL_657108\|20 | ···················· | ···················· | ···················· | ···················· | ···················· | ···················· |
| S | 3200 hCoV-19/England/CAMC-B5459A/2020\|EPI_ISL_658381\|20 | ···················· | ···················· | ···················· | ···················· | ···················· | ···················· |
| S | 3201 hCoV-19/England/MILK-B5A6EC/2020\|EPI_ISL_658384\|20 | ···················· | ···················· | ···················· | ···················· | ···················· | ···················· |
| S | 3202 hCoV-19/England/MILK-B5A479/2020\|EPI_ISL_658385\|20 | ···················· | ···················· | ···················· | ···················· | ···················· | ···················· |
| S | 3203 hCoV-19/England/MILK-B5A646/2020\|EPI_ISL_658386\|20 | ···················· | ···················· | ···················· | ···················· | ···················· | ···················· |
| S | 3204 hCoV-19/England/CAMC-B54785/2020\|EPI_ISL_658387\|20 | ···················· | ···················· | ···················· | ···················· | ······T············· | ···················· |
| S | 3205 hCoV-19/England/CAMC-B516B8/2020\|EPI_ISL_658402\|20 | ···················· | ···················· | ···················· | ···················· | ···················· | ···················· |
| S | 3206 hCoV-19/England/MILK-B5603D/2020\|EPI_ISL_658403\|20 | ···················· | ···················· | ···················· | ···················· | ···················· | ···················· |
| S | 3207 hCoV-19/England/QEUH-B3F322/2020\|EPI_ISL_658405\|20 | ···················· | ·A·················· | ···················· | ···················· | ···················· | ···················· |
| S | 3208 hCoV-19/England/QEUH-B411F6/2020\|EPI_ISL_658406\|20 | ···················· | ···················· | ···················· | ···················· | ···················· | ···················· |
| S | 3209 hCoV-19/England/QEUH-B41897/2020\|EPI_ISL_658407\|20 | ···················· | ···················· | ···················· | ···················· | ···················· | ···················· |
| S | 3210 hCoV-19/England/QEUH-B3F44D/2020\|EPI_ISL_658408\|20 | ···················· | ···················· | ···················· | ···················· | ···················· | ···················· |
| S | 3211 hCoV-19/England/QEUH-B41008/2020\|EPI_ISL_658411\|20 | ···················· | ·A·················· | ···················· | ···················· | ···················· | ··A················· |
| S | 3212 hCoV-19/England/QEUH-B4178B/2020\|EPI_ISL_658413\|20 | ···················· | ···················· | ···················· | ···················· | ···················· | ···················· |
| S | 3213 hCoV-19/England/QEUH-B3F94E/2020\|EPI_ISL_658414\|20 | ···················· | ···················· | ···················· | ···················· | ···················· | ···················· |

| | | | | | | | |
|---|---|---|---|---|---|---|---|
| S | 3214 hCoV-19/England/QEUH-B419EF/2020\|EPI_ISL_658415\|20 | ···················· | ···················· | ···················· | ···················· | ···················· | ···················· |
| S | 3215 hCoV-19/England/QEUH-B41563/2020\|EPI_ISL_658416\|20 | ···················· | ···················· | ···················· | ···················· | ···················· | ···················· |
| S | 3216 hCoV-19/England/QEUH-B41114/2020\|EPI_ISL_658417\|20 | ···················· | ···················· | ···················· | ···················· | ···················· | ···················· |
| S | 3217 hCoV-19/England/QEUH-B419FE/2020\|EPI_ISL_658418\|20 | ···················· | ···················· | ···················· | ···················· | ···················· | ···················· |
| S | 3218 hCoV-19/England/QEUH-B3F805/2020\|EPI_ISL_658419\|20 | ···················· | ···················· | ···················· | ···················· | ···················· | ···················· |
| S | 3219 hCoV-19/England/QEUH-B3F586/2020\|EPI_ISL_658420\|20 | ···················· | ···················· | ···················· | ···················· | ···················· | ···················· |
| S | 3220 hCoV-19/England/QEUH-B412C6/2020\|EPI_ISL_658421\|20 | ···················· | ···················· | ···················· | ···················· | ···················· | ···················· |
| S | 3221 hCoV-19/England/QEUH-B41590/2020\|EPI_ISL_658422\|20 | ···················· | ···················· | ···················· | ···················· | ···················· | ···················· |
| S | 3222 hCoV-19/England/QEUH-B4131E/2020\|EPI_ISL_658423\|20 | ···················· | ···················· | ···················· | ···················· | ···················· | ···················· |
| S | 3223 hCoV-19/England/QEUH-B4126C/2020\|EPI_ISL_658424\|20 | ···················· | ···················· | ···················· | ···················· | ···················· | ···················· |
| S | 3224 hCoV-19/England/QEUH-B3F2CB/2020\|EPI_ISL_658425\|20 | ···················· | ···A················ | ···················· | ···················· | ···················· | ···················· |
| S | 3225 hCoV-19/England/QEUH-B3FDBB/2020\|EPI_ISL_658426\|20 | ···················· | ···················· | ···················· | ···················· | ···················· | ···················· |
| S | 3226 hCoV-19/England/QEUH-B413C3/2020\|EPI_ISL_658427\|20 | ···················· | ···················· | ···················· | ···················· | ···················· | ···················· |
| S | 3227 hCoV-19/England/QEUH-B3FB48/2020\|EPI_ISL_658428\|20 | ···················· | ···················· | ···················· | ···················· | ···················· | ···················· |
| S | 3228 hCoV-19/England/QEUH-B415BE/2020\|EPI_ISL_658430\|20 | ···················· | ···················· | ···················· | ···················· | ···················· | ···················· |
| S | 3229 hCoV-19/England/QEUH-B4134B/2020\|EPI_ISL_658431\|20 | ···················· | ···················· | ···················· | ···················· | ···················· | ···················· |
| S | 3230 hCoV-19/England/QEUH-B41123/2020\|EPI_ISL_658432\|20 | ···················· | ···················· | ···················· | ···················· | ···················· | ···················· |
| S | 3231 hCoV-19/England/QEUH-B3F7CC/2020\|EPI_ISL_658433\|20 | ···················· | ···················· | ···················· | ···················· | ···················· | ···················· |
| S | 3232 hCoV-19/England/QEUH-B41B52/2020\|EPI_ISL_658434\|20 | ···················· | ···················· | ···················· | ···················· | ···················· | ···················· |
| S | 3233 hCoV-19/England/QEUH-B415CD/2020\|EPI_ISL_658435\|20 | ···················· | ···················· | ···················· | ···················· | ···················· | ···················· |
| S | 3234 hCoV-19/England/QEUH-B3F726/2020\|EPI_ISL_658436\|20 | ···················· | ···················· | ···················· | ···················· | ···················· | ···················· |
| S | 3235 hCoV-19/England/QEUH-B3F902/2020\|EPI_ISL_658437\|20 | ···················· | ···················· | ···················· | ···················· | ···················· | ···················· |
| S | 3236 hCoV-19/England/QEUH-B413D2/2020\|EPI_ISL_658438\|20 | ···················· | ···················· | ···················· | ···················· | ···················· | ···················· |
| S | 3237 hCoV-19/England/QEUH-B40FFD/2020\|EPI_ISL_658439\|20 | ···················· | ·A·················· | ···················· | ···················· | ···················· | ···A················ |
| S | 3238 hCoV-19/England/CAMC-B357E4/2020\|EPI_ISL_659701\|20 | ···················· | ···················· | ···················· | ···················· | ···················· | ···················· |
| S | 3239 hCoV-19/England/CAMC-B3290F/2020\|EPI_ISL_659702\|20 | ···················· | ···················· | ···················· | ···················· | ···················· | ···················· |
| S | 3240 hCoV-19/England/CAMC-B34017/2020\|EPI_ISL_659706\|20 | ···················· | ···················· | ···················· | ···················· | ···················· | ···················· |
| S | 3241 hCoV-19/England/CAMC-B34888/2020\|EPI_ISL_659707\|20 | ···················· | ···················· | ···················· | ···················· | ···················· | ···················· |
| S | 3242 hCoV-19/England/CAMC-B34123/2020\|EPI_ISL_659708\|20 | ···················· | ···················· | ···················· | ···················· | ···················· | ···················· |
| S | 3243 hCoV-19/England/CAMC-B32C9D/2020\|EPI_ISL_659713\|20 | ···················· | ···················· | ···················· | ···················· | ···················· | ···················· |
| S | 3244 hCoV-19/England/CAMC-B32B54/2020\|EPI_ISL_659714\|20 | ···················· | ···················· | ···················· | ···················· | ···················· | ···················· |
| S | 3245 hCoV-19/England/CAMC-B3416F/2020\|EPI_ISL_659716\|20 | ···················· | ···················· | ···················· | ···················· | ···················· | ···················· |
| S | 3246 hCoV-19/England/CAMC-B34BAD/2020\|EPI_ISL_659717\|20 | ···················· | ···················· | ···················· | ···················· | ···················· | ···················· |
| S | 3247 hCoV-19/England/CAMC-B32B18/2020\|EPI_ISL_659718\|20 | ···················· | ···················· | ···················· | ···················· | ···················· | ···················· |
| S | 3248 hCoV-19/England/CAMC-B3468E/2020\|EPI_ISL_659719\|20 | ···················· | ···················· | ···················· | ···················· | ···················· | ···················· |
| S | 3249 hCoV-19/England/CAMC-B32CD9/2020\|EPI_ISL_659720\|20 | ···················· | ···················· | ···················· | ···················· | ···················· | ···················· |
| S | 3250 hCoV-19/England/CAMC-B34396/2020\|EPI_ISL_659721\|20 | ···················· | ···················· | ···················· | ···················· | ···················· | ···················· |
| S | 3251 hCoV-19/England/CAMC-B33FDF/2020\|EPI_ISL_659722\|20 | ···················· | ···················· | ···················· | ···················· | ···················· | ···················· |
| S | 3252 hCoV-19/England/CAMC-B32E00/2020\|EPI_ISL_659723\|20 | ···················· | ···················· | ···················· | ···················· | ···················· | ···················· |
| S | 3253 hCoV-19/England/CAMC-B3582D/2020\|EPI_ISL_659725\|20 | ···················· | ·A·················· | ···················· | ···················· | ···················· | ···················· |
| S | 3254 hCoV-19/England/CAMC-B34202/2020\|EPI_ISL_659726\|20 | ···················· | ···················· | ···················· | ···················· | ···················· | ···················· |
| S | 3255 hCoV-19/England/CAMC-B345CD/2020\|EPI_ISL_659727\|20 | ···················· | ···················· | ···················· | ···················· | ···················· | ···················· |
| S | 3256 hCoV-19/England/CAMC-B347E5/2020\|EPI_ISL_659728\|20 | ···················· | ···················· | ···················· | ···················· | ···················· | ···················· |
| S | 3257 hCoV-19/England/CAMC-B340DB/2020\|EPI_ISL_659731\|20 | ···················· | ·A·················· | ···················· | ···················· | ···················· | ···A················ |
| S | 3258 hCoV-19/England/CAMC-B34493/2020\|EPI_ISL_659732\|20 | ···················· | ···················· | ···················· | ···················· | ···················· | ···················· |
| S | 3259 hCoV-19/England/CAMC-B34633/2020\|EPI_ISL_659733\|20 | ···················· | ···················· | ···················· | ···················· | ···················· | ···················· |
| S | 3260 hCoV-19/England/CAMC-B34994/2020\|EPI_ISL_659735\|20 | ···················· | ···················· | ···················· | ···················· | ···················· | ···················· |
| S | 3261 hCoV-19/England/CAMC-B34518/2020\|EPI_ISL_659736\|20 | ···················· | ···················· | ···················· | ···················· | ···················· | ···················· |
| S | 3262 hCoV-19/England/CAMC-B341D8/2020\|EPI_ISL_659738\|20 | ···················· | ···················· | ···················· | ···················· | ···················· | ···················· |
| S | 3263 hCoV-19/England/CAMC-B349FE/2020\|EPI_ISL_659740\|20 | ···················· | ·A·················· | ···················· | ···················· | ···················· | ···A················ |
| S | 3264 hCoV-19/England/CAMC-B345EB/2020\|EPI_ISL_659741\|20 | ···················· | ·A·················· | ···················· | ···················· | ···················· | ···A················ |
| S | 3265 hCoV-19/England/CAMC-B34AA0/2020\|EPI_ISL_659743\|20 | ···················· | ···················· | ···················· | ···················· | ···················· | ···················· |
| S | 3266 hCoV-19/England/QEUH-B2CE63/2020\|EPI_ISL_659744\|20 | ···················· | ···················· | ···················· | ···················· | ···················· | ···················· |
| S | 3267 hCoV-19/England/QEUH-B2FC93/2020\|EPI_ISL_659746\|20 | ···················· | ···················· | ···················· | ···················· | ···················· | ···················· |
| S | 3268 hCoV-19/England/QEUH-B2F932/2020\|EPI_ISL_659747\|20 | ···················· | ···················· | ···················· | ···················· | ···················· | ···················· |
| S | 3269 hCoV-19/England/QEUH-B2E818/2020\|EPI_ISL_659751\|20 | ···················· | ···················· | ···················· | ···················· | ···················· | ···················· |
| S | 3270 hCoV-19/England/QEUH-B2DF8D/2020\|EPI_ISL_659752\|20 | ···················· | ···················· | ···················· | ···················· | ···················· | ···················· |
| S | 3271 hCoV-19/England/QEUH-B2D275/2020\|EPI_ISL_659753\|20 | ···················· | ···················· | ···················· | ···················· | ···················· | ···················· |
| S | 3272 hCoV-19/England/QEUH-B2E960/2020\|EPI_ISL_659754\|20 | ···················· | ···················· | ···················· | ···················· | ···················· | ···················· |
| S | 3273 hCoV-19/England/QEUH-B2D5E5/2020\|EPI_ISL_659756\|20 | ···················· | ···················· | ···················· | ···················· | ···················· | ···················· |
| S | 3274 hCoV-19/England/QEUH-B2F316/2020\|EPI_ISL_659758\|20 | ···················· | ···················· | ···················· | ···················· | ···················· | ···················· |
| S | 3275 hCoV-19/England/QEUH-B2CE45/2020\|EPI_ISL_659759\|20 | ···················· | ···················· | ···················· | ···················· | ···················· | ···················· |
| S | 3276 hCoV-19/England/QEUH-B30321/2020\|EPI_ISL_659760\|20 | ···················· | ···················· | ···················· | ···················· | ···················· | ···················· |
| S | 3277 hCoV-19/England/QEUH-B2D327/2020\|EPI_ISL_659761\|20 | ···················· | ···················· | ···················· | ···················· | ···················· | ···················· |
| S | 3278 hCoV-19/England/QEUH-B2CE27/2020\|EPI_ISL_659762\|20 | ···················· | ···················· | ···················· | ···················· | ···················· | ···················· |
| S | 3279 hCoV-19/England/QEUH-B2DF23/2020\|EPI_ISL_659763\|20 | ···················· | ···················· | ···················· | ···················· | ···················· | ···················· |
| S | 3280 hCoV-19/England/QEUH-B2D63D/2020\|EPI_ISL_659765\|20 | ···················· | ···················· | ···················· | ···················· | ···················· | ···················· |
| S | 3281 hCoV-19/England/QEUH-B30B1A/2020\|EPI_ISL_659766\|20 | ···················· | ···················· | ···················· | ···················· | ···················· | ···················· |
| S | 3282 hCoV-19/England/QEUH-B2CEDC/2020\|EPI_ISL_659767\|20 | ···················· | ···················· | ···················· | ···················· | ···················· | ···················· |
| S | 3283 hCoV-19/France/OCC-SC509/2020\|EPI_ISL_660700\|2020- | ···················· | ···················· | ···················· | ···················· | ···················· | ···················· |
| S | 3284 hCoV-19/France/OCC-SC510/2020\|EPI_ISL_660701\|2020- | ···················· | ···················· | ···················· | ···················· | ···················· | ···················· |
| S | 3285 hCoV-19/France/OCC-SC511/2020\|EPI_ISL_660702\|2020- | ···················· | ···················· | ···················· | ···················· | ···················· | ···················· |
| S | 3286 hCoV-19/France/OCC-SC512/2020\|EPI_ISL_660703\|2020- | ···················· | ···················· | ···················· | ···················· | ···················· | ···················· |
| S | 3288 hCoV-19/France/OCC-SC517/2020\|EPI_ISL_660705\|2020- | ···················· | ···················· | ···················· | ···················· | ···················· | ···················· |
| S | 3289 hCoV-19/France/OCC-SC518/2020\|EPI_ISL_660706\|2020- | ···················· | ···················· | ···················· | ···················· | ···················· | ···················· |
| S | 3290 hCoV-19/France/OCC-SC520/2020\|EPI_ISL_660707\|2020- | ···················· | ···················· | ···················· | ···················· | ···················· | ···················· |
| S | 3291 hCoV-19/France/OCC-SC521/2020\|EPI_ISL_660708\|2020- | ···················· | ···················· | ···················· | ···················· | ···················· | ···················· |
| S | 3292 hCoV-19/France/OCC-SC522/2020\|EPI_ISL_660709\|2020- | ···················· | ···················· | ···················· | ···················· | ···················· | ···················· |
| S | 3293 hCoV-19/France/OCC-SC525/2020\|EPI_ISL_660710\|2020- | ···················· | ···················· | ···················· | ···················· | ···················· | ···················· |
| S | 3294 hCoV-19/France/OCC-SC526/2020\|EPI_ISL_660711\|2020- | ···················· | ···················· | ···················· | ···················· | ···················· | ···················· |

| | | | | | | | |
|---|---|---|---|---|---|---|---|
| S | 3295 hCoV-19/France/OCC-SC528/2020\|EPI_ISL_660712\|2020- | ···················· | ·········A·········· | ····················· | ···················· | ···················· | ···················· |
| S | 3296 hCoV-19/France/OCC-SC536/2020\|EPI_ISL_660713\|2020- | ···················· | ···················· | ····················· | ···················· | ···················· | ···················· |
| S | 3297 hCoV-19/France/OCC-SC537/2020\|EPI_ISL_660714\|2020- | ···················· | ···················· | ····················· | ···················· | ···················· | ···················· |
| S | 3298 hCoV-19/France/OCC-SC538/2020\|EPI_ISL_660715\|2020- | ···················· | ···················· | ····················· | ···················· | ···················· | ···················· |
| S | 3299 hCoV-19/France/OCC-SC539/2020\|EPI_ISL_660716\|2020- | ···················· | ···················· | ····················· | ···················· | ···················· | ···················· |
| S | 3300 hCoV-19/France/OCC-SC541/2020\|EPI_ISL_660717\|2020- | ···················· | ···················· | ····················· | ···················· | ···················· | ···················· |
| S | 3301 hCoV-19/France/OCC-SC542/2020\|EPI_ISL_660718\|2020- | ···················· | ···················· | ····················· | ···················· | ···················· | ···················· |
| S | 3302 hCoV-19/France/OCC-SC544/2020\|EPI_ISL_660719\|2020- | ···················· | ···················· | ····················· | ···················· | ···················· | ···················· |
| S | 3303 hCoV-19/France/OCC-SC548/2020\|EPI_ISL_660720\|2020- | ···················· | ···················· | ····················· | ···················· | ···················· | ···················· |
| S | 3304 hCoV-19/France/OCC-SC550/2020\|EPI_ISL_660721\|2020- | ···················· | ·········A·········· | ····················· | ···················· | ···················· | ···················· |
| S | 3305 hCoV-19/France/OCC-SC551/2020\|EPI_ISL_660722\|2020- | ···················· | ·········A·········· | ····················· | ···················· | ···················· | ···················· |
| S | 3306 hCoV-19/France/OCC-SC552/2020\|EPI_ISL_660723\|2020- | ···················· | ·········A·········· | ····················· | ···················· | ···················· | ···················· |
| S | 3307 hCoV-19/France/OCC-SC555/2020\|EPI_ISL_660724\|2020- | ···················· | ···················· | ····················· | ···················· | ···················· | ···················· |
| S | 3308 hCoV-19/France/OCC-SC556/2020\|EPI_ISL_660725\|2020- | ···················· | ···················· | ····················· | ···················· | ···················· | ···················· |
| S | 3309 hCoV-19/France/OCC-SC558/2020\|EPI_ISL_660726\|2020- | ···················· | ···················· | ····················· | ···················· | ···················· | ···················· |
| S | 3310 hCoV-19/France/OCC-SC560/2020\|EPI_ISL_660727\|2020- | ···················· | ···················· | ····················· | ···················· | ···················· | ···················· |
| S | 3311 hCoV-19/France/OCC-SC561/2020\|EPI_ISL_660728\|2020- | ···················· | ···················· | ····················· | ···················· | ···················· | ···················· |
| S | 3312 hCoV-19/France/OCC-SC563/2020\|EPI_ISL_660729\|2020- | ···················· | ···················· | ····················· | ···················· | ···················· | ···················· |
| S | 3313 hCoV-19/France/OCC-SC564/2020\|EPI_ISL_660730\|2020- | ···················· | ·········A·········· | ····················· | ···················· | ···················· | ···················· |
| S | 3314 hCoV-19/France/OCC-SC565/2020\|EPI_ISL_660731\|2020- | ···················· | ···················· | ····················· | ···················· | ···················· | ···················· |
| S | 3315 hCoV-19/France/PAC-SC439/2020\|EPI_ISL_660732\|2020- | ···················· | ···················· | ····················· | ···················· | ···················· | ···················· |
| S | 3316 hCoV-19/France/PAC-SC440/2020\|EPI_ISL_660733\|2020- | ···················· | ···················· | ····················· | ···················· | ···················· | ···················· |
| S | 3317 hCoV-19/France/PAC-SC441/2020\|EPI_ISL_660734\|2020- | ···················· | ···················· | ····················· | ···················· | ···················· | ···················· |
| S | 3318 hCoV-19/France/PAC-SC444/2020\|EPI_ISL_660735\|2020- | ···················· | ···················· | ····················· | ···················· | ···················· | ···················· |
| S | 3319 hCoV-19/France/PAC-SC447/2020\|EPI_ISL_660736\|2020- | ···················· | ···················· | ····················· | ···················· | ···················· | ···················· |
| S | 3320 hCoV-19/France/PAC-SC450/2020\|EPI_ISL_660737\|2020- | ···················· | ···················· | ····················· | ···················· | ···················· | ···················· |
| S | 3321 hCoV-19/France/PAC-SC453/2020\|EPI_ISL_660738\|2020- | ···················· | ···················· | ····················· | ···················· | ···················· | ···················· |
| S | 3322 hCoV-19/France/PAC-SC454/2020\|EPI_ISL_660739\|2020- | ···················· | ···················· | ····················· | ···················· | ···················· | ···················· |
| S | 3323 hCoV-19/France/PAC-SC455/2020\|EPI_ISL_660740\|2020- | ···················· | ···················· | ····················· | ···················· | ···················· | ···················· |
| S | 3324 hCoV-19/France/PAC-SC459/2020\|EPI_ISL_660741\|2020- | ···················· | ···················· | ····················· | ···················· | ···················· | ···················· |
| S | 3325 hCoV-19/England/204291949/2020\|EPI_ISL_660743\|2020 | ···················· | ···················· | ····················· | ···················· | ···················· | ···················· |
| S | 3326 hCoV-19/England/204291952/2020\|EPI_ISL_660744\|2020 | ···················· | ···················· | ····················· | ···················· | ···················· | ···················· |
| S | 3327 hCoV-19/England/204291969/2020\|EPI_ISL_660745\|2020 | ···················· | ·A·················· | ····················· | ···················· | ···················· | ··A················· |
| S | 3328 hCoV-19/England/204380343/2020\|EPI_ISL_660747\|2020 | ···················· | ···················· | ····················· | ···················· | ···················· | ···················· |
| S | 3329 hCoV-19/England/MILK-B05862/2020\|EPI_ISL_652200\|20 | ···················· | ···················· | ····················· | ···················· | ···················· | ···················· |
| S | 3330 hCoV-19/England/MILK-B05880/2020\|EPI_ISL_652201\|20 | ···················· | ···················· | ····················· | ···················· | ···················· | ···················· |
| S | 3331 hCoV-19/Wales/MILK-B058CC/2020\|EPI_ISL_652202\|2020 | ···················· | ···················· | ····················· | ···················· | ···················· | ···················· |
| S | 3332 hCoV-19/England/MILK-B058DB/2020\|EPI_ISL_652203\|20 | ···················· | ···················· | ····················· | ···················· | ···················· | ···················· |
| S | 3333 hCoV-19/Wales/MILK-B058EA/2020\|EPI_ISL_652204\|2020 | ···················· | ···················· | ····················· | ···················· | ···················· | ···················· |
| S | 3334 hCoV-19/England/MILK-B058F9/2020\|EPI_ISL_652205\|20 | ···················· | ···················· | ····················· | ···················· | ···················· | ···················· |
| S | 3335 hCoV-19/England/MILK-B05914/2020\|EPI_ISL_652206\|20 | ···················· | ···················· | ····················· | ···················· | ···················· | ···················· |
| S | 3336 hCoV-19/England/MILK-B05941/2020\|EPI_ISL_652207\|20 | ···················· | ···················· | ····················· | ···················· | ···················· | ···················· |
| S | 3337 hCoV-19/England/MILK-B05950/2020\|EPI_ISL_652208\|20 | ···················· | ···················· | ····················· | ···················· | ···················· | ···················· |
| S | 3338 hCoV-19/England/MILK-B0598D/2020\|EPI_ISL_652209\|20 | ···················· | ···················· | ····················· | ···················· | ·····T·············· | ···················· |
| S | 3339 hCoV-19/England/MILK-B0599C/2020\|EPI_ISL_652210\|20 | ···················· | ···················· | ····················· | ···················· | ···················· | ···················· |
| S | 3340 hCoV-19/England/MILK-B059C9/2020\|EPI_ISL_652211\|20 | ···················· | ···················· | ····················· | ···················· | ···················· | ···················· |
| S | 3341 hCoV-19/England/MILK-B059D8/2020\|EPI_ISL_652212\|20 | ···················· | ···················· | ····················· | ···················· | ···················· | ···················· |
| S | 3342 hCoV-19/England/MILK-B059E7/2020\|EPI_ISL_652213\|20 | ···················· | ···················· | ····················· | ···················· | ···················· | ···················· |
| S | 3343 hCoV-19/USA/LA-EVTL819/2020\|EPI_ISL_653510\|2020-07 | ···················· | ···················· | ····················· | ···················· | ···················· | ···················· |
| S | 3344 hCoV-19/USA/LA-EVTL820/2020\|EPI_ISL_653511\|2020-07 | ···················· | ···················· | ····················· | ···················· | ···················· | ···················· |
| S | 3345 hCoV-19/USA/LA-EVTL832/2020\|EPI_ISL_653523\|2020-07 | ···················· | ···················· | ····················· | ···················· | ···················· | ···················· |
| S | 3346 hCoV-19/USA/LA-EVTL833/2020\|EPI_ISL_653524\|2020-07 | ···················· | ···················· | ····················· | ···················· | ···················· | ···················· |
| S | 3347 hCoV-19/USA/LA-EVTL834/2020\|EPI_ISL_653525\|2020-07 | ···················· | ···················· | ····················· | ···················· | ···················· | ···················· |
| S | 3348 hCoV-19/USA/LA-EVTL835/2020\|EPI_ISL_653526\|2020-07 | ···················· | ···················· | ····················· | ···················· | ···················· | ···················· |
| S | 3349 hCoV-19/USA/LA-EVTL836/2020\|EPI_ISL_653527\|2020-07 | ···················· | ···················· | ····················· | ···················· | ···················· | ···················· |
| S | 3350 hCoV-19/USA/LA-EVTL837/2020\|EPI_ISL_653528\|2020-07 | ···················· | ···················· | ····················· | ···················· | ···················· | ···················· |
| S | 3351 hCoV-19/USA/LA-EVTL838/2020\|EPI_ISL_653529\|2020-07 | ···················· | ···················· | ····················· | ···················· | ···················· | ···················· |
| S | 3352 hCoV-19/USA/LA-EVTL839/2020\|EPI_ISL_653530\|2020-07 | ···················· | ···················· | ····················· | ···················· | ···················· | ···················· |
| S | 3353 hCoV-19/USA/LA-EVTL840/2020\|EPI_ISL_653531\|2020-07 | ···················· | ···················· | ····················· | ···················· | ···················· | ···················· |
| S | 3354 hCoV-19/USA/LA-EVTL841/2020\|EPI_ISL_653532\|2020-07 | ···················· | ···················· | ····················· | ···················· | ···················· | ···················· |
| S | 3355 hCoV-19/USA/LA-EVTL842/2020\|EPI_ISL_653533\|2020-07 | ···················· | ·A·················· | ····················· | ···················· | ···················· | ···················· |
| S | 3356 hCoV-19/USA/LA-EVTL843/2020\|EPI_ISL_653534\|2020-07 | ···················· | ···················· | ····················· | ···················· | ···················· | ···················· |
| S | 3357 hCoV-19/USA/LA-EVTL844/2020\|EPI_ISL_653535\|2020-07 | ···················· | ···················· | ····················· | ···················· | ···················· | ···················· |
| S | 3358 hCoV-19/USA/LA-EVTL845/2020\|EPI_ISL_653536\|2020-07 | ···················· | ···················· | ····················· | ···················· | ···················· | ···················· |
| S | 3359 hCoV-19/USA/LA-EVTL846/2020\|EPI_ISL_653537\|2020-07 | ···················· | ···················· | ····················· | ···················· | ···················· | ···················· |
| S | 3360 hCoV-19/USA/LA-EVTL847/2020\|EPI_ISL_653538\|2020-07 | ···················· | ···················· | ····················· | ···················· | ···················· | ···················· |
| S | 3361 hCoV-19/USA/LA-EVTL848/2020\|EPI_ISL_653539\|2020-07 | ···················· | ···················· | ····················· | ···················· | ···················· | ···················· |
| S | 3362 hCoV-19/USA/LA-EVTL849/2020\|EPI_ISL_653540\|2020-07 | ···················· | ···················· | ····················· | ···················· | ···················· | ···················· |
| S | 3363 hCoV-19/USA/LA-EVTL850/2020\|EPI_ISL_653541\|2020-07 | ···················· | ·A·················· | ····················· | ···················· | ···················· | ···················· |
| S | 3364 hCoV-19/USA/LA-EVTL851/2020\|EPI_ISL_653542\|2020-07 | ···················· | ···················· | ····················· | ···················· | ···················· | ···················· |
| S | 3365 hCoV-19/USA/LA-EVTL852/2020\|EPI_ISL_653543\|2020-07 | ···················· | ···················· | ····················· | ···················· | ···················· | ···················· |
| S | 3366 hCoV-19/USA/LA-EVTL853/2020\|EPI_ISL_653544\|2020-07 | ···················· | ···················· | ····················· | ···················· | ···················· | ···················· |
| S | 3367 hCoV-19/Vietnam/PIHCM-14/2020\|EPI_ISL_654866\|2020- | ···················· | ···················· | ····················· | ···················· | ···················· | ···················· |
| S | 3368 hCoV-19/Vietnam/PIHCM-304/2020\|EPI_ISL_654867\|2020 | ···················· | ···················· | ····················· | ···················· | ···················· | ···················· |
| S | 3369 hCoV-19/Vietnam/PIHCM-321/2020\|EPI_ISL_654868\|2020 | ···················· | ···················· | ····················· | ···················· | ···················· | ···················· |
| S | 3370 hCoV-19/Vietnam/PIHCM-322/2020\|EPI_ISL_654869\|2020 | ···················· | ···················· | ····················· | ···················· | ···················· | ···················· |
| S | 3371 hCoV-19/Vietnam/PIHCM-323/2020\|EPI_ISL_654870\|2020 | ···················· | ···················· | ····················· | ···················· | ···················· | ···················· |
| S | 3372 hCoV-19/Vietnam/PIHCM-372/2020\|EPI_ISL_654872\|2020 | ···················· | ···················· | ····················· | ···················· | ···················· | ···················· |
| S | 3373 hCoV-19/Vietnam/PIHCM-406/2020\|EPI_ISL_654873\|2020 | ···················· | ···················· | ····················· | ···················· | ···················· | ···················· |
| S | 3374 hCoV-19/Vietnam/PIHCM-459/2020\|EPI_ISL_654874\|2020 | ···················· | ···················· | ····················· | ···················· | ···················· | ···················· |

S 3375 hCoV-19/Vietnam/PIHCM-461/2020|EPI_ISL_654875|2020 ···················· ···················· ····················· ···················· ···················· ····················
S 3376 hCoV-19/England/CAMC-B52347/2020|EPI_ISL_657000|20 ···················· ···················· ····················· ···················· ···················· ····················
S 3377 hCoV-19/England/CAMC-B541F0/2020|EPI_ISL_657001|20 ···················· ···················· ····················· ···················· ···················· ····················
S 3378 hCoV-19/England/CAMC-B5349E/2020|EPI_ISL_657002|20 ···················· ·A·················· ····················· ···················· ···················· ···A················
S 3379 hCoV-19/England/MILK-B56CEE/2020|EPI_ISL_657003|20 ···················· ···················· ····················· ···················· ···················· ····················
S 3380 hCoV-19/England/MILK-B56756/2020|EPI_ISL_657004|20 ···················· ·A·················· ····················· ···················· ···················· ···A················
S 3381 hCoV-19/England/CAMC-B52769/2020|EPI_ISL_657006|20 ···················· ···················· ····················· ···················· ···················· ····················
S 3382 hCoV-19/England/CAMC-B5365C/2020|EPI_ISL_657007|20 ···················· ···················· ····················· ···················· ···················· ····················
S 3383 hCoV-19/England/MILK-B56BF0/2020|EPI_ISL_657008|20 ···················· ·A·················· ····················· ···················· ···················· ···A················
S 3384 hCoV-19/England/CAMC-B5406C/2020|EPI_ISL_657009|20 ···················· ···················· ····················· ···················· ···················· ····················
S 3385 hCoV-19/England/CAMC-B524AE/2020|EPI_ISL_657010|20 ···················· ···················· ····················· ···················· ···················· ····················
S 3386 hCoV-19/England/MILK-B56CDF/2020|EPI_ISL_657011|20 ···················· ···················· ····················· ···················· ···················· ····················
S 3387 hCoV-19/England/CAMC-B538ED/2020|EPI_ISL_657013|20 ···················· ···················· ····················· ···················· ···················· ····················
S 3388 hCoV-19/England/MILK-B56826/2020|EPI_ISL_657014|20 ···················· ···················· ················G···· ···················· ···················· ····················
S 3389 hCoV-19/England/CAMC-B5223B/2020|EPI_ISL_657015|20 ···················· ···················· ····················· ···················· ···················· ····················
S 3390 hCoV-19/England/MILK-B56B3C/2020|EPI_ISL_657016|20 ···················· ···················· ····················· ···················· ···················· ····················
S 3391 hCoV-19/England/MILK-B56905/2020|EPI_ISL_657017|20 ···················· ···················· ····················· ···················· ···················· ····················
S 3392 hCoV-19/England/CAMC-B5405D/2020|EPI_ISL_657018|20 ···················· ···················· ····················· ···················· ···················· ····················
S 3393 hCoV-19/England/CAMC-B533A0/2020|EPI_ISL_657019|20 ···················· ·A·················· ····················· ···················· ···················· ····················
S 3394 hCoV-19/England/CAMC-B537A4/2020|EPI_ISL_657020|20 ···················· ···················· ····················· ···················· ···················· ····················
S 3395 hCoV-19/England/MILK-B56BE1/2020|EPI_ISL_657021|20 ···················· ···················· ····················· ···················· ···················· ····················
S 3396 hCoV-19/England/CAMC-B53786/2020|EPI_ISL_657022|20 ···················· ···················· ····················· ···················· ···················· ····················
S 3397 hCoV-19/England/MILK-B56DDC/2020|EPI_ISL_657023|20 ···················· ···················· ····················· ···················· ···················· ····················
S 3398 hCoV-19/England/CAMC-B5356E/2020|EPI_ISL_657025|20 ···················· ···················· ····················· ···················· ···················· ····················
S 3399 hCoV-19/England/MILK-B56C1B/2020|EPI_ISL_657026|20 ···················· ···················· ····················· ···················· ···················· ····················
S 3400 hCoV-19/England/QEUH-B3FFE2/2020|EPI_ISL_657027|20 ···················· ···················· ····················· ···················· ···················· ····················
S 3401 hCoV-19/England/QEUH-B422B6/2020|EPI_ISL_657028|20 ···················· ···················· ····················· ···················· ···················· ····················
S 3402 hCoV-19/England/QEUH-B4078C/2020|EPI_ISL_657029|20 ···················· ···················· ····················· ···················· ···················· ····················
S 3403 hCoV-19/England/QEUH-B3FF4C/2020|EPI_ISL_657030|20 ···················· ···················· ····················· ···················· ···················· ····················
S 3404 hCoV-19/England/QEUH-B432C4/2020|EPI_ISL_657031|20 ···················· ···················· ····················· ···················· ···················· ····················
S 3405 hCoV-19/England/QEUH-B43473/2020|EPI_ISL_657032|20 ···················· ···················· ····················· ···················· ···················· ····················
S 3406 hCoV-19/England/QEUH-B4241A/2020|EPI_ISL_657033|20 ···················· ···················· ····················· ···················· ···················· ····················
S 3407 hCoV-19/England/QEUH-B42650/2020|EPI_ISL_657034|20 ···················· ···················· ····················· ···················· ···················· ····················
S 3408 hCoV-19/England/QEUH-B420DA/2020|EPI_ISL_657035|20 ···················· ···················· ····················· ···················· ···················· ····················
S 3409 hCoV-19/England/QEUH-B4259F/2020|EPI_ISL_657036|20 ···················· ···················· ····················· ···················· ···················· ····················
S 3410 hCoV-19/England/QEUH-B432F1/2020|EPI_ISL_657037|20 ···················· ···················· ····················· ···················· ···················· ····················
S 3411 hCoV-19/England/QEUH-B42535/2020|EPI_ISL_657038|20 ···················· ···················· ····················· ···················· ···················· ····················
S 3412 hCoV-19/England/QEUH-B406CB/2020|EPI_ISL_657039|20 ···················· ···················· ····················· ···················· ···················· ····················
S 3413 hCoV-19/England/QEUH-B407E6/2020|EPI_ISL_657040|20 ···················· ···················· ····················· ···················· ···················· ····················
S 3414 hCoV-19/England/QEUH-B42298/2020|EPI_ISL_657041|20 ···················· ···················· ····················· ···················· ···················· ····················
S 3415 hCoV-19/England/QEUH-B4025E/2020|EPI_ISL_657042|20 ···················· ···················· ····················· ···················· ···················· ····················
S 3416 hCoV-19/England/QEUH-B4227A/2020|EPI_ISL_657043|20 ···················· ···················· ····················· ···················· ···················· ····················
S 3417 hCoV-19/England/QEUH-B42580/2020|EPI_ISL_657044|20 ···················· ···················· ····················· ···················· ···················· ····················
S 3418 hCoV-19/England/QEUH-B405FB/2020|EPI_ISL_657045|20 ···················· ···················· ····················· ···················· ···················· ····················
S 3419 hCoV-19/England/QEUH-B3FF5B/2020|EPI_ISL_657046|20 ···················· ···················· ····················· ···················· ···················· ····················
S 3420 hCoV-19/England/QEUH-B424CF/2020|EPI_ISL_657047|20 ···················· ···················· ····················· ···················· ···················· ····················
S 3421 hCoV-19/England/QEUH-B407D7/2020|EPI_ISL_657048|20 ···················· ···················· ····················· ···················· ···················· ····················
S 3422 hCoV-19/England/QEUH-B426BA/2020|EPI_ISL_657049|20 ···················· ···················· ····················· ···················· ···················· ····················
S 3423 hCoV-19/England/QEUH-B436C8/2020|EPI_ISL_657050|20 ···················· ···················· ····················· ···················· ···················· ····················
S 3424 hCoV-19/England/QEUH-B40151/2020|EPI_ISL_657051|20 ···················· ···················· ····················· ···················· ···················· ····················
S 3425 hCoV-19/England/QEUH-B40546/2020|EPI_ISL_657052|20 ···················· ···················· ····················· ···················· ···················· ····················
S 3426 hCoV-19/England/QEUH-B3FF1F/2020|EPI_ISL_657057|20 ···················· ···················· ····················· ···················· ···················· ····················
S 3427 hCoV-19/England/QEUH-B3FDF7/2020|EPI_ISL_657059|20 ···················· ···················· ····················· ···················· ···················· ····················
S 3428 hCoV-19/England/QEUH-B4019D/2020|EPI_ISL_657060|20 ···················· ···················· ····················· ···················· ···················· ····················
S 3429 hCoV-19/England/QEUH-B4090E/2020|EPI_ISL_657061|20 ···················· ···················· ····················· ···················· ···················· ····················
S 3430 hCoV-19/England/QEUH-B43437/2020|EPI_ISL_657062|20 ···················· ···················· ····················· ···················· ···················· ····················
S 3431 hCoV-19/England/QEUH-B40018/2020|EPI_ISL_657063|20 ···················· ···················· ····················· ···················· ···················· ····················
S 3432 hCoV-19/England/QEUH-B3FF6A/2020|EPI_ISL_657064|20 ···················· ···················· ····················· ···················· ···················· ····················
S 3433 hCoV-19/England/QEUH-B4092C/2020|EPI_ISL_657065|20 ···················· ···················· ····················· ···················· ···················· ····················
S 3434 hCoV-19/England/QEUH-B401CA/2020|EPI_ISL_657066|20 ···················· ···················· ····················· ···················· ···················· ····················
S 3435 hCoV-19/England/QEUH-B42623/2020|EPI_ISL_657067|20 ···················· ·A·················· ···A················· ···················· ···················· ···A················
S 3436 hCoV-19/England/CAMC-B518A3/2020|EPI_ISL_658320|20 ···················· ···················· ····················· ···················· ···················· ····················
S 3437 hCoV-19/England/CAMC-B546A6/2020|EPI_ISL_658321|20 ···················· ·A·················· ····················· ···················· ···················· ···A················
S 3438 hCoV-19/England/MILK-B5A400/2020|EPI_ISL_658322|20 ···················· ···················· ····················· ···················· ···················· ····················
S 3439 hCoV-19/England/CAMC-B51797/2020|EPI_ISL_658323|20 ···················· ···················· ····················· ···················· ···················· ····················
S 3440 hCoV-19/England/MILK-B55DCE/2020|EPI_ISL_658324|20 ···················· ···················· ····················· ···················· ···················· ····················
S 3441 hCoV-19/England/MILK-B5A7AD/2020|EPI_ISL_658325|20 ···················· ···················· ····················· ···················· ···················· ····················
S 3442 hCoV-19/England/CAMC-B54758/2020|EPI_ISL_658326|20 ···················· ···················· ····················· ···················· ···················· ····················
S 3443 hCoV-19/England/CAMC-B51858/2020|EPI_ISL_658327|20 ···················· ···················· ····················· ···················· ···················· ····················
S 3444 hCoV-19/England/MILK-B5604C/2020|EPI_ISL_658328|20 ···················· ···················· ····················· ···················· ···················· ····················
S 3445 hCoV-19/England/MILK-B56079/2020|EPI_ISL_658329|20 ···················· ···················· ····················· ···················· ···················· ····················
S 3446 hCoV-19/England/CAMC-B51A16/2020|EPI_ISL_658330|20 ···················· ···················· ····················· ···················· ···················· ····················
S 3447 hCoV-19/England/CAMC-B51B40/2020|EPI_ISL_658331|20 ···················· ···················· ····················· ···················· ···················· ····················
S 3448 hCoV-19/England/CAMC-B546D3/2020|EPI_ISL_658332|20 ···················· ···················· ····················· ···················· ···················· ····················
S 3449 hCoV-19/England/MILK-B55E61/2020|EPI_ISL_658333|20 ···················· ···················· ····················· ···················· ···················· ····················
S 3450 hCoV-19/England/CAMC-B54882/2020|EPI_ISL_658334|20 ···················· ···················· ····················· ···················· ···················· ····················
S 3451 hCoV-19/England/MILK-B5A770/2020|EPI_ISL_658336|20 ···················· ···················· ····················· ···················· ···················· ····················
S 3452 hCoV-19/England/CAMC-B54891/2020|EPI_ISL_658337|20 ···················· ···················· ····················· ···················· ···················· ····················
S 3453 hCoV-19/England/MILK-B55FD7/2020|EPI_ISL_658338|20 ···················· ···················· ····················· ···················· ···················· ····················
S 3454 hCoV-19/England/MILK-B5A51C/2020|EPI_ISL_658339|20 ···················· ···················· ····················· ···················· ···················· ····················

| | | | | | | | | |
|---|---|---|---|---|---|---|---|---|
| S | 3455 | hCoV-19/England/CAMC-B547C1/2020\|EPI_ISL_658340\|20 | ···················· | ···················· | ···················· | ···················· | ···················· | ···················· |
| S | 3456 | hCoV-19/England/MILK-B561D0/2020\|EPI_ISL_658342\|20 | ···················· | ···················· | ···················· | ···················· | ···················· | ···················· |
| S | 3457 | hCoV-19/England/MILK-B55E25/2020\|EPI_ISL_658344\|20 | ···················· | ···················· | ···················· | ···················· | ···················· | ···················· |
| S | 3458 | hCoV-19/England/MILK-B55D19/2020\|EPI_ISL_658346\|20 | ···················· | ···················· | ···················· | ···················· | ···················· | ···················· |
| S | 3459 | hCoV-19/England/MILK-B55C2B/2020\|EPI_ISL_658347\|20 | ···················· | ···················· | ···················· | ···················· | ···················· | ···················· |
| S | 3460 | hCoV-19/England/CAMC-B5466A/2020\|EPI_ISL_658348\|20 | ···················· | ···················· | ···················· | ···················· | ···················· | ···················· |
| S | 3461 | hCoV-19/England/MILK-B55D55/2020\|EPI_ISL_658349\|20 | ···················· | ···················· | ···················· | ···················· | ···················· | ···················· |
| S | 3462 | hCoV-19/England/CAMC-B51BAA/2020\|EPI_ISL_658352\|20 | ···················· | ···················· | ···················· | ···················· | ···················· | ···················· |
| S | 3463 | hCoV-19/England/MILK-B5601F/2020\|EPI_ISL_658354\|20 | ···················· | ···················· | ···················· | ···················· | ···················· | ···················· |
| S | 3464 | hCoV-19/England/CAMC-B549AD/2020\|EPI_ISL_658356\|20 | ···················· | ···················· | ···················· | ···················· | ···················· | ···················· |
| S | 3465 | hCoV-19/England/MILK-B5A558/2020\|EPI_ISL_658357\|20 | ···················· | ···················· | ···················· | ···················· | ···················· | ···················· |
| S | 3466 | hCoV-19/England/MILK-B55CD0/2020\|EPI_ISL_658358\|20 | ···················· | ···················· | ···················· | ···················· | ···················· | ···················· |
| S | 3467 | hCoV-19/England/MILK-B55D73/2020\|EPI_ISL_658359\|20 | ···················· | ···················· | ···················· | ···················· | ···················· | ···A················ |
| S | 3468 | hCoV-19/England/CAMC-B51788/2020\|EPI_ISL_658360\|20 | ···················· | ···················· | ···················· | ···················· | ···················· | ···················· |
| S | 3469 | hCoV-19/England/CAMC-B54916/2020\|EPI_ISL_658361\|20 | ···················· | ···················· | ···················· | ···················· | ···················· | ···················· |
| S | 3470 | hCoV-19/England/CAMC-B515BB/2020\|EPI_ISL_658362\|20 | ···················· | ·A······A··········· | ···················· | ···················· | ···················· | ···················· |
| S | 3471 | hCoV-19/England/CAMC-B519BF/2020\|EPI_ISL_658364\|20 | ···················· | ···················· | ···················· | ···················· | ···················· | ···················· |
| S | 3472 | hCoV-19/England/MILK-B56000/2020\|EPI_ISL_658365\|20 | ···················· | ···················· | ···················· | ···················· | ···················· | ···················· |
| S | 3473 | hCoV-19/England/MILK-B55F9B/2020\|EPI_ISL_658369\|20 | ···················· | ···················· | ···················· | ···················· | ···················· | ···················· |
| S | 3474 | hCoV-19/England/CAMC-B5171F/2020\|EPI_ISL_658370\|20 | ···················· | ···················· | ···················· | ···················· | ···················· | ···················· |
| S | 3475 | hCoV-19/England/CAMC-B54819/2020\|EPI_ISL_658371\|20 | ···················· | ···················· | ···················· | ···················· | ···················· | ···················· |
| S | 3476 | hCoV-19/England/MILK-B55FF5/2020\|EPI_ISL_658372\|20 | ···················· | ···················· | ···················· | ···················· | ···················· | ···················· |
| S | 3477 | hCoV-19/England/MILK-B5A78F/2020\|EPI_ISL_658373\|20 | ···················· | ···················· | ···················· | ···················· | ···················· | ···················· |
| S | 3478 | hCoV-19/England/MILK-B55CA3/2020\|EPI_ISL_658374\|20 | ···················· | ···················· | ···················· | ···················· | ···················· | ···················· |
| S | 3479 | hCoV-19/England/MILK-B5A7BC/2020\|EPI_ISL_658375\|20 | ···················· | ···················· | ···················· | ···················· | ···················· | ···················· |
| S | 3480 | hCoV-19/England/MILK-B5A330/2020\|EPI_ISL_658377\|20 | ···················· | ···················· | ···················· | ···················· | ···················· | ···················· |
| S | 3481 | hCoV-19/England/MILK-B56158/2020\|EPI_ISL_658378\|20 | ···················· | ···················· | ···················· | ···················· | ···················· | ···················· |
| S | 3482 | hCoV-19/England/MILK-B5A2E8/2020\|EPI_ISL_658379\|20 | ···················· | ···················· | ···················· | ···················· | ···················· | ···················· |
| S | 3483 | hCoV-19/England/CAMC-B51867/2020\|EPI_ISL_658383\|20 | ···················· | ···················· | ···················· | ···················· | ···················· | ···················· |
| S | 3484 | hCoV-19/England/MILK-B5A6CE/2020\|EPI_ISL_658389\|20 | ···················· | ···················· | ···················· | ···················· | ···················· | ···················· |
| S | 3485 | hCoV-19/England/CAMC-B545B8/2020\|EPI_ISL_658390\|20 | ···················· | ···················· | ···················· | ···················· | ···················· | ···················· |
| S | 3486 | hCoV-19/England/MILK-B55FB9/2020\|EPI_ISL_658391\|20 | ···················· | ···················· | ···················· | ···················· | ···················· | ···················· |
| S | 3487 | hCoV-19/England/MILK-B5A6DD/2020\|EPI_ISL_658392\|20 | ···················· | ···················· | ···················· | ···················· | ···················· | ···················· |
| S | 3488 | hCoV-19/England/MILK-B5A6BF/2020\|EPI_ISL_658393\|20 | ···················· | ···················· | ···················· | ···················· | ···················· | ···················· |
| S | 3489 | hCoV-19/England/MILK-B55E34/2020\|EPI_ISL_658395\|20 | ···················· | ···················· | ···················· | ···················· | ···················· | ···················· |
| S | 3490 | hCoV-19/England/CAMC-B54688/2020\|EPI_ISL_658396\|20 | ···················· | ·A·················· | ···················· | ···················· | ···················· | ···A················ |
| S | 3491 | hCoV-19/England/CAMC-B546F1/2020\|EPI_ISL_658397\|20 | ···················· | ···················· | ···················· | ···················· | ···················· | ···················· |
| S | 3492 | hCoV-19/England/MILK-B5A36D/2020\|EPI_ISL_658399\|20 | ···················· | ···················· | ···················· | ···················· | ···················· | ···················· |
| S | 3493 | hCoV-19/England/CAMC-B33FC0/2020\|EPI_ISL_659650\|20 | ···················· | ···················· | ···················· | ···················· | ···················· | ···················· |
| S | 3494 | hCoV-19/England/CAMC-B357D5/2020\|EPI_ISL_659651\|20 | ···················· | ···················· | ···················· | ···················· | ···················· | ···················· |
| S | 3495 | hCoV-19/England/CAMC-B3287B/2020\|EPI_ISL_659652\|20 | ···················· | ···················· | ···················· | ···················· | ···················· | ···················· |
| S | 3496 | hCoV-19/England/CAMC-B32C60/2020\|EPI_ISL_659653\|20 | ···················· | ···················· | ···················· | ···················· | ······T············· | ···············A···· |
| S | 3497 | hCoV-19/England/CAMC-B34A55/2020\|EPI_ISL_659654\|20 | ···················· | ···················· | ···················· | ···················· | ···················· | ···················· |
| S | 3498 | hCoV-19/England/CAMC-B34A37/2020\|EPI_ISL_659656\|20 | ···················· | ···················· | ···················· | ···················· | ······T············· | ···················· |
| S | 3499 | hCoV-19/England/CAMC-B35483/2020\|EPI_ISL_659657\|20 | ···················· | ···················· | ···················· | ···················· | ···················· | ···················· |
| S | 3500 | hCoV-19/England/CAMC-B35711/2020\|EPI_ISL_659658\|20 | ···················· | ···················· | ···················· | ···················· | ···················· | ···················· |
| S | 3501 | hCoV-19/England/CAMC-B348A6/2020\|EPI_ISL_659659\|20 | ···················· | ···················· | ···················· | ···················· | ···················· | ···················· |
| S | 3502 | hCoV-19/England/CAMC-B34132/2020\|EPI_ISL_659660\|20 | ···················· | ···················· | ···················· | ···················· | ···················· | ···················· |
| S | 3503 | hCoV-19/England/CAMC-B34624/2020\|EPI_ISL_659663\|20 | ···················· | ···················· | ···················· | ···················· | ···················· | ···················· |
| S | 3504 | hCoV-19/England/CAMC-B35140/2020\|EPI_ISL_659664\|20 | ···················· | ···················· | ···················· | ···················· | ···················· | ···················· |
| S | 3505 | hCoV-19/England/CAMC-B34660/2020\|EPI_ISL_659665\|20 | ···················· | ·A·················· | ···················· | ···················· | ···················· | ···A················ |
| S | 3506 | hCoV-19/England/CAMC-B342D5/2020\|EPI_ISL_659667\|20 | ···················· | ···················· | ···················· | ···················· | ···················· | ···················· |
| S | 3507 | hCoV-19/England/CAMC-B354ED/2020\|EPI_ISL_659668\|20 | ···················· | ···················· | ···················· | ···················· | ···················· | ···················· |
| S | 3508 | hCoV-19/England/CAMC-B3584B/2020\|EPI_ISL_659669\|20 | ···················· | ···················· | ···················· | ···················· | ···················· | ···················· |
| S | 3509 | hCoV-19/England/CAMC-B346D9/2020\|EPI_ISL_659670\|20 | ···················· | ···················· | ···················· | ···················· | ···················· | ···················· |
| S | 3510 | hCoV-19/England/CAMC-B32AA2/2020\|EPI_ISL_659671\|20 | ···················· | ···················· | ···················· | ···················· | ···················· | ···················· |
| S | 3511 | hCoV-19/England/CAMC-B34985/2020\|EPI_ISL_659672\|20 | ···················· | ···················· | ···················· | ···················· | ···················· | ···················· |
| S | 3512 | hCoV-19/England/CAMC-B340BD/2020\|EPI_ISL_659673\|20 | ···················· | ···················· | ···················· | ···················· | ···················· | ···················· |
| S | 3513 | hCoV-19/England/CAMC-B356D8/2020\|EPI_ISL_659675\|20 | ···················· | ·A·················· | ···················· | ···················· | ···················· | ···················· |
| S | 3514 | hCoV-19/England/CAMC-B347F4/2020\|EPI_ISL_659676\|20 | ···················· | ···················· | ···················· | ···················· | ······T············· | ···················· |
| S | 3515 | hCoV-19/England/CAMC-B35052/2020\|EPI_ISL_659677\|20 | ···················· | ···················· | ···················· | ···················· | ···················· | ···················· |
| S | 3516 | hCoV-19/England/CAMC-B329D2/2020\|EPI_ISL_659680\|20 | ···················· | ···················· | ···················· | ···················· | ···················· | ···················· |
| S | 3517 | hCoV-19/England/CAMC-B34150/2020\|EPI_ISL_659681\|20 | ···················· | ···················· | ···················· | ···················· | ···················· | ···················· |
| S | 3518 | hCoV-19/England/CAMC-B3474F/2020\|EPI_ISL_659682\|20 | ···················· | ···················· | ···················· | ···················· | ···················· | ···················· |
| S | 3519 | hCoV-19/England/CAMC-B3578A/2020\|EPI_ISL_659684\|20 | ···················· | ···················· | ···················· | ···················· | ···················· | ···················· |
| S | 3520 | hCoV-19/England/CAMC-B3430F/2020\|EPI_ISL_659685\|20 | ···················· | ···················· | ···················· | ···················· | ···················· | ···················· |
| S | 3521 | hCoV-19/England/CAMC-B355CC/2020\|EPI_ISL_659686\|20 | ···················· | ···················· | ···················· | ···················· | ···················· | ···················· |
| S | 3522 | hCoV-19/England/CAMC-B3475E/2020\|EPI_ISL_659688\|20 | ···················· | ···················· | ···················· | ···················· | ···················· | ···················· |
| S | 3523 | hCoV-19/England/CAMC-B356AB/2020\|EPI_ISL_659692\|20 | ···················· | ·A·················· | ···················· | ···················· | ···················· | ···A················ |
| S | 3524 | hCoV-19/England/CAMC-B34044/2020\|EPI_ISL_659693\|20 | ···················· | ···················· | ···················· | ···················· | ···················· | ···················· |
| S | 3525 | hCoV-19/England/CAMC-B32B72/2020\|EPI_ISL_659694\|20 | ···················· | ···················· | ···················· | ···················· | ···················· | ···················· |
| S | 3526 | hCoV-19/England/CAMC-B3279C/2020\|EPI_ISL_659695\|20 | ···················· | ···················· | ···················· | ···················· | ···················· | ···················· |
| S | 3527 | hCoV-19/England/CAMC-B32A75/2020\|EPI_ISL_659697\|20 | ···················· | ···················· | ···················· | ···················· | ···················· | ···················· |
| S | 3528 | hCoV-19/South Africa/KRISP-K004601/2020\|EPI_ISL_66 | ···················· | ···················· | ···················· | ···················· | ···················· | ···················· |
| S | 3529 | hCoV-19/South Africa/KRISP-K004602/2020\|EPI_ISL_66 | ···················· | ···················· | ···················· | ···················· | ···················· | ···················· |
| S | 3530 | hCoV-19/South Africa/KRISP-K004606/2020\|EPI_ISL_66 | ···················· | ···················· | ···················· | ···················· | ···················· | ···················· |
| S | 3531 | hCoV-19/South Africa/KRISP-K004608/2020\|EPI_ISL_66 | ···················· | ···················· | ···················· | ···················· | ···················· | ···················· |
| S | 3532 | hCoV-19/South Africa/KRISP-K004609/2020\|EPI_ISL_66 | ···················· | ···················· | ···················· | ···················· | ···················· | ···················· |
| S | 3533 | hCoV-19/South Africa/KRISP-K004610/2020\|EPI_ISL_66 | ···················· | ···················· | ···················· | ···················· | ···················· | ···················· |
| S | 3534 | hCoV-19/South Africa/KRISP-K004613/2020\|EPI_ISL_66 | ···················· | ·A·················· | ···················· | ···················· | ···················· | ···················· |

| | | | | | | | | |
|---|---|---|---|---|---|---|---|---|
| S | 3535 hCoV-19/South Africa/KRISP-K004616/2020\|EPI_ISL_66 | .................... | .................... | .................... | .................... | .................... | .................... | |
| S | 3536 hCoV-19/South Africa/KRISP-K004621/2020\|EPI_ISL_66 | .................... | .................... | ..T................. | .................... | .................... | .................... | |
| S | 3537 hCoV-19/South Africa/KRISP-K004622/2020\|EPI_ISL_66 | .................... | .................... | .................... | .................... | .................... | .................... | |
| S | 3538 hCoV-19/South Africa/KRISP-K004625/2020\|EPI_ISL_66 | .................... | .................... | .................... | .................... | .................... | .................... | |
| S | 3539 hCoV-19/South Africa/KRISP-K004626/2020\|EPI_ISL_66 | .................... | .................... | .................... | .................... | .................... | .................... | |
| S | 3540 hCoV-19/South Africa/KRISP-K004627/2020\|EPI_ISL_66 | .................... | .................... | .................... | .................... | .................... | .................... | |
| S | 3541 hCoV-19/South Africa/KRISP-K004628/2020\|EPI_ISL_66 | .................... | .................... | .................... | .................... | .................... | .................... | |
| S | 3542 hCoV-19/South Africa/KRISP-K004629/2020\|EPI_ISL_66 | .................... | .A.................. | .................... | .................... | .................... | .................... | |
| S | 3543 hCoV-19/South Africa/KRISP-K004633/2020\|EPI_ISL_66 | .................... | .A.................. | .................... | .................... | .................... | .................... | |
| S | 3544 hCoV-19/South Africa/KRISP-K004634/2020\|EPI_ISL_66 | .................... | .................... | .................... | .................... | .................... | .................... | |
| S | 3545 hCoV-19/South Africa/KRISP-K004635/2020\|EPI_ISL_66 | .................... | .................... | .................... | .................... | .................... | .................... | |
| S | 3546 hCoV-19/South Africa/KRISP-K004636/2020\|EPI_ISL_66 | .................... | .................... | ..T................. | .................... | .................... | .................... | |
| S | 3547 hCoV-19/South Africa/KRISP-K004639/2020\|EPI_ISL_66 | .................... | .................... | .................... | .................... | .................... | .................... | |
| S | 3548 hCoV-19/South Africa/KRISP-K004640/2020\|EPI_ISL_66 | .................... | .................... | .................... | .................... | .................... | .................... | |
| S | 3549 hCoV-19/South Africa/KRISP-K004642/2020\|EPI_ISL_66 | .................... | .................... | .................... | .................... | .................... | .................... | |
| S | 3550 hCoV-19/South Africa/KRISP-K004647/2020\|EPI_ISL_66 | .................... | .A.................. | .................... | .................... | .................... | .................... | |
| S | 3551 hCoV-19/South Africa/KRISP-K004649/2020\|EPI_ISL_66 | .................... | .................... | .................... | .................... | .................... | .................... | |
| S | 3552 hCoV-19/South Africa/KRISP-K004650/2020\|EPI_ISL_66 | .................... | .................... | .................... | .................... | .................... | .................... | |
| S | 3553 hCoV-19/France/OCC-SC425/2020\|EPI_ISL_660665\|2020- | .................... | .................... | .................... | .................... | .................... | .................... | |
| S | 3554 hCoV-19/France/OCC-SC426/2020\|EPI_ISL_660666\|2020- | .................... | .................... | .................... | .................... | .................... | .................... | |
| S | 3555 hCoV-19/France/OCC-SC428/2020\|EPI_ISL_660667\|2020- | .................... | .................... | .................... | .................... | .................... | .................... | |
| S | 3556 hCoV-19/France/OCC-SC429/2020\|EPI_ISL_660668\|2020- | .................... | .................... | .................... | .................... | .................... | .................... | |
| S | 3557 hCoV-19/France/OCC-SC433/2020\|EPI_ISL_660670\|2020- | .................... | .................... | .................... | .................... | .................... | .................... | |
| S | 3558 hCoV-19/France/OCC-SC434/2020\|EPI_ISL_660671\|2020- | .................... | .................... | .................... | .................... | .................... | .................... | |
| S | 3559 hCoV-19/France/OCC-SC464/2020\|EPI_ISL_660672\|2020- | .................... | .................... | .................... | .................... | .................... | .................... | |
| S | 3560 hCoV-19/France/OCC-SC469/2020\|EPI_ISL_660673\|2020- | .................... | .................... | .................... | .................... | .................... | .................... | |
| S | 3561 hCoV-19/France/OCC-SC470/2020\|EPI_ISL_660674\|2020- | .................... | .................... | .................... | .................... | .................... | .................... | |
| S | 3562 hCoV-19/France/OCC-SC473/2020\|EPI_ISL_660675\|2020- | .................... | .................... | .................... | .................... | .................... | .................... | |
| S | 3563 hCoV-19/France/OCC-SC475/2020\|EPI_ISL_660676\|2020- | .................... | .................... | .................... | .................... | .................... | .................... | |
| S | 3564 hCoV-19/France/OCC-SC476/2020\|EPI_ISL_660677\|2020- | .................... | .................... | .................... | .................... | .................... | .................... | |
| S | 3565 hCoV-19/France/OCC-SC477/2020\|EPI_ISL_660678\|2020- | .................... | .................... | .................... | .................... | .................... | .................... | |
| S | 3566 hCoV-19/France/OCC-SC478/2020\|EPI_ISL_660679\|2020- | .................... | ...................A | .................... | .................... | .................... | .................... | 1 |
| S | 3567 hCoV-19/France/OCC-SC479/2020\|EPI_ISL_660680\|2020- | .................... | .................... | .................... | .................... | .................... | .................... | |
| S | 3568 hCoV-19/France/OCC-SC480/2020\|EPI_ISL_660681\|2020- | .................... | ...................A | .................... | .................... | .................... | .................... | 1 |
| S | 3569 hCoV-19/France/OCC-SC481/2020\|EPI_ISL_660682\|2020- | .................... | .................... | .................... | .................... | .................... | .................... | |
| S | 3570 hCoV-19/France/OCC-SC482/2020\|EPI_ISL_660683\|2020- | .................... | .................... | .................... | .................... | .................... | .................... | |
| S | 3571 hCoV-19/France/OCC-SC484/2020\|EPI_ISL_660684\|2020- | .................... | .................... | .................... | .................... | .................... | .................... | |
| S | 3572 hCoV-19/France/OCC-SC485/2020\|EPI_ISL_660685\|2020- | .................... | ..........A......... | .................... | .................... | .................... | .................... | |
| S | 3573 hCoV-19/France/OCC-SC488/2020\|EPI_ISL_660686\|2020- | .................... | .................... | .................... | .................... | .................... | .................... | |
| S | 3574 hCoV-19/France/OCC-SC490/2020\|EPI_ISL_660687\|2020- | .................... | .................... | .................... | .................... | .................... | .................... | |
| S | 3575 hCoV-19/France/OCC-SC492/2020\|EPI_ISL_660688\|2020- | .................... | .................... | .................... | .................... | .................... | .................... | |
| S | 3576 hCoV-19/France/OCC-SC494/2020\|EPI_ISL_660689\|2020- | .................... | .................... | .................... | .................... | .................... | .................... | |
| S | 3577 hCoV-19/France/OCC-SC495/2020\|EPI_ISL_660690\|2020- | .................... | .................... | .................... | .................... | .................... | .................... | |
| S | 3578 hCoV-19/France/OCC-SC497/2020\|EPI_ISL_660691\|2020- | .................... | .................... | .................... | .................... | .................... | .................... | |
| S | 3579 hCoV-19/France/OCC-SC498/2020\|EPI_ISL_660692\|2020- | .................... | ...................A | .................... | .................... | .................... | .................... | 1 |
| S | 3580 hCoV-19/France/OCC-SC499/2020\|EPI_ISL_660693\|2020- | .................... | .................... | .................... | .................... | .................... | .................... | |
| S | 3581 hCoV-19/France/OCC-SC500/2020\|EPI_ISL_660694\|2020- | .................... | .................... | .................... | .................... | .................... | .................... | |
| S | 3582 hCoV-19/France/OCC-SC501/2020\|EPI_ISL_660695\|2020- | .................... | .................... | .................... | .................... | .................... | .................... | |
| S | 3583 hCoV-19/France/OCC-SC502/2020\|EPI_ISL_660696\|2020- | .................... | .................... | .................... | .................... | .................... | .................... | |
| S | 3584 hCoV-19/France/OCC-SC505/2020\|EPI_ISL_660697\|2020- | .................... | .................... | .................... | .................... | .................... | .................... | |
| S | 3585 hCoV-19/England/LIVE-DB64D3/2020\|EPI_ISL_652100\|20 | .................... | .................... | .................... | .................... | .................... | .................... | |
| S | 3586 hCoV-19/England/LIVE-DB651C/2020\|EPI_ISL_652101\|20 | .................... | .................... | .................... | .................... | .................... | .................... | |
| S | 3587 hCoV-19/England/LIVE-DB652B/2020\|EPI_ISL_652102\|20 | .................... | .................... | .................... | .................... | .................... | .................... | |
| S | 3588 hCoV-19/England/MILK-A23EC7/2020\|EPI_ISL_652103\|20 | .................... | .................... | .................... | .................... | .................... | .................... | |
| S | 3589 hCoV-19/Wales/MILK-A23F1F/2020\|EPI_ISL_652104\|2020 | .................... | .................... | .................... | .................... | .................... | .................... | |
| S | 3590 hCoV-19/Wales/MILK-A23F3D/2020\|EPI_ISL_652105\|2020 | .................... | .................... | .................... | .................... | .................... | .................... | |
| S | 3591 hCoV-19/England/MILK-A23FB5/2020\|EPI_ISL_652106\|20 | .................... | .A.................. | .................... | .................... | .................... | .................... | |
| S | 3592 hCoV-19/England/MILK-A24066/2020\|EPI_ISL_652107\|20 | .................... | .A.................. | .................... | .................... | .................... | .................... | |
| S | 3593 hCoV-19/England/MILK-A240EE/2020\|EPI_ISL_652108\|20 | .................... | .................... | .................... | .................... | ......T............. | .................... | |
| S | 3594 hCoV-19/England/MILK-A240FD/2020\|EPI_ISL_652109\|20 | .................... | .................... | .................... | .................... | ......T............. | .................... | |
| S | 3595 hCoV-19/England/MILK-A24118/2020\|EPI_ISL_652110\|20 | .................... | .................... | .................... | .................... | .................... | .................... | |
| S | 3596 hCoV-19/England/MILK-A8EA02/2020\|EPI_ISL_652111\|20 | .................... | .................... | .................... | .................... | .................... | .................... | |
| S | 3597 hCoV-19/England/MILK-A8EAC6/2020\|EPI_ISL_652112\|20 | .................... | .................... | .................... | .................... | .................... | .................... | |
| S | 3598 hCoV-19/England/MILK-A8EC84/2020\|EPI_ISL_652113\|20 | .................... | .................... | .................... | .................... | .................... | .................... | |
| S | 3599 hCoV-19/England/MILK-A8EEE8/2020\|EPI_ISL_652114\|20 | .................... | .................... | .................... | .................... | ......T............. | .................... | |
| S | 3600 hCoV-19/England/MILK-A8EF6D/2020\|EPI_ISL_652115\|20 | .................... | .................... | .................... | .................... | .................... | .................... | |
| S | 3601 hCoV-19/England/MILK-A8F05A/2020\|EPI_ISL_652116\|20 | .................... | .................... | .................... | .................... | .................... | .................... | |
| S | 3602 hCoV-19/England/MILK-ACF8ED/2020\|EPI_ISL_652117\|20 | .................... | .A.......A.......... | .................... | .................... | .................... | .................... | |
| S | 3603 hCoV-19/England/MILK-ACFA8D/2020\|EPI_ISL_652118\|20 | .................... | .................... | .................... | .................... | .................... | .................... | |
| S | 3604 hCoV-19/England/MILK-ACFA9C/2020\|EPI_ISL_652119\|20 | .................... | .................... | .................... | .................... | .................... | .................... | |
| S | 3605 hCoV-19/England/MILK-ACFAAB/2020\|EPI_ISL_652120\|20 | .................... | .................... | .................... | .................... | .................... | .................... | |
| S | 3606 hCoV-19/England/MILK-ACFAC9/2020\|EPI_ISL_652121\|20 | .................... | .................... | .................... | .................... | .................... | .................... | |
| S | 3607 hCoV-19/England/MILK-ACFAD8/2020\|EPI_ISL_652122\|20 | .................... | .................... | .................... | .................... | .................... | .................... | |
| S | 3608 hCoV-19/England/MILK-ACFB4E/2020\|EPI_ISL_652123\|20 | .................... | .................... | .................... | .................... | .................... | .................... | |
| S | 3609 hCoV-19/England/MILK-ACFB6C/2020\|EPI_ISL_652124\|20 | .................... | .A.................. | .................... | .................... | .................... | ...A................ | |
| S | 3610 hCoV-19/England/MILK-ACFB7B/2020\|EPI_ISL_652125\|20 | .................... | .................... | .................... | .................... | .................... | .................... | |
| S | 3611 hCoV-19/England/MILK-ACFB99/2020\|EPI_ISL_652126\|20 | .................... | .................... | .................... | .................... | .................... | .................... | |
| S | 3612 hCoV-19/England/MILK-ACFC1E/2020\|EPI_ISL_652127\|20 | .................... | .................... | .................... | .................... | .................... | .................... | |
| S | 3613 hCoV-19/England/MILK-ACFC3C/2020\|EPI_ISL_652128\|20 | .................... | .................... | .................... | .................... | ......T............. | .................... | |
| S | 3614 hCoV-19/England/MILK-ACFC4B/2020\|EPI_ISL_652129\|20 | .................... | .................... | .................... | .................... | .................... | .................... | |

| | | | | | | | |
|---|---|---|---|---|---|---|---|
| S | 3615 hCoV-19/England/MILK-ACFC5A/2020\|EPI_ISL_652130\|20 | .................... | .................... | ..................... | .................... | .................... | .................... |
| S | 3616 hCoV-19/England/MILK-ACFCA5/2020\|EPI_ISL_652131\|20 | .................... | .................... | ..................... | .................... | .................... | .................... |
| S | 3617 hCoV-19/England/MILK-ACFCB4/2020\|EPI_ISL_652132\|20 | .................... | .A.................. | ..................... | .................... | .................... | ..A................. |
| S | 3618 hCoV-19/England/MILK-ACFCC3/2020\|EPI_ISL_652133\|20 | .................... | .................... | ..................... | .................... | .................... | .................... |
| S | 3619 hCoV-19/England/MILK-ACFCD2/2020\|EPI_ISL_652134\|20 | .................... | .................... | ..................... | .................... | .................... | .................... |
| S | 3620 hCoV-19/England/MILK-ACFD2A/2020\|EPI_ISL_652135\|20 | .................... | .................... | ..................... | .................... | .................... | .................... |
| S | 3621 hCoV-19/England/MILK-ACFD48/2020\|EPI_ISL_652136\|20 | .................... | .................... | ..................... | .................... | .................... | .................... |
| S | 3622 hCoV-19/England/MILK-ACFD57/2020\|EPI_ISL_652137\|20 | .................... | .................... | ..................... | .................... | .................... | .................... |
| S | 3623 hCoV-19/England/MILK-ACFDC0/2020\|EPI_ISL_652138\|20 | .................... | .................... | ..................... | .................... | .................... | .................... |
| S | 3624 hCoV-19/England/MILK-ACFE36/2020\|EPI_ISL_652139\|20 | .................... | .................... | ..................... | .................... | .................... | .................... |
| S | 3625 hCoV-19/England/MILK-ACFE63/2020\|EPI_ISL_652140\|20 | .................... | .................... | ..................... | .................... | .................... | .................... |
| S | 3626 hCoV-19/England/MILK-ACFE81/2020\|EPI_ISL_652141\|20 | .................... | .................... | ..................... | .................... | .................... | .................... |
| S | 3627 hCoV-19/England/MILK-ACFE90/2020\|EPI_ISL_652142\|20 | .................... | .................... | ..................... | .................... | .................... | .................... |
| S | 3628 hCoV-19/England/MILK-ACFEBE/2020\|EPI_ISL_652143\|20 | .................... | .................... | ..................... | .................... | .................... | .................... |
| S | 3629 hCoV-19/England/MILK-ACFEEB/2020\|EPI_ISL_652144\|20 | .................... | .................... | ..................... | .................... | ......T............. | .................... |
| S | 3630 hCoV-19/England/MILK-ACFEFA/2020\|EPI_ISL_652145\|20 | .................... | .................... | ..................... | .................... | .................... | .................... |
| S | 3631 hCoV-19/England/MILK-ACFF06/2020\|EPI_ISL_652146\|20 | .................... | .................... | ..................... | .................... | ......T............. | .................... |
| S | 3632 hCoV-19/England/MILK-AD003C/2020\|EPI_ISL_652147\|20 | .................... | .................... | ..................... | .................... | .................... | .................... |
| S | 3633 hCoV-19/England/MILK-AD004B/2020\|EPI_ISL_652148\|20 | .................... | .................... | ..................... | .................... | .................... | .................... |
| S | 3634 hCoV-19/England/MILK-AD0078/2020\|EPI_ISL_652149\|20 | .................... | .................... | ..................... | .................... | .................... | .................... |
| S | 3635 hCoV-19/England/MILK-AD0087/2020\|EPI_ISL_652150\|20 | .................... | .................... | ..................... | .................... | .................... | .................... |
| S | 3636 hCoV-19/England/MILK-B02DFD/2020\|EPI_ISL_652151\|20 | .................... | .................... | ..................... | .................... | .................... | .................... |
| S | 3637 hCoV-19/Wales/MILK-B02E45/2020\|EPI_ISL_652152\|2020 | .................... | .................... | ..................... | .................... | .................... | .................... |
| S | 3638 hCoV-19/Wales/MILK-B02E63/2020\|EPI_ISL_652153\|2020 | .................... | .................... | ..................... | .................... | .................... | .................... |
| S | 3639 hCoV-19/England/MILK-B02E72/2020\|EPI_ISL_652154\|20 | .................... | .................... | ..................... | .................... | .................... | .................... |
| S | 3640 hCoV-19/England/MILK-B02EEB/2020\|EPI_ISL_652155\|20 | .................... | .................... | ..................... | .................... | .................... | .................... |
| S | 3641 hCoV-19/England/MILK-B0303F/2020\|EPI_ISL_652156\|20 | .................... | .................... | ..................... | .................... | .................... | .................... |
| S | 3642 hCoV-19/England/MILK-B044C9/2020\|EPI_ISL_652157\|20 | .................... | .................... | ..................... | .................... | .................... | .................... |
| S | 3643 hCoV-19/England/MILK-B04502/2020\|EPI_ISL_652158\|20 | .................... | .................... | ..................... | .................... | .................... | .................... |
| S | 3644 hCoV-19/England/MILK-B04511/2020\|EPI_ISL_652159\|20 | .................... | .................... | ..................... | .................... | .................... | .................... |
| S | 3645 hCoV-19/England/MILK-B04520/2020\|EPI_ISL_652160\|20 | .................... | .................... | ..................... | .................... | .................... | .................... |
| S | 3646 hCoV-19/England/MILK-B0453F/2020\|EPI_ISL_652161\|20 | .................... | .................... | ..................... | .................... | .................... | .................... |
| S | 3647 hCoV-19/England/MILK-B045A8/2020\|EPI_ISL_652162\|20 | .................... | .................... | ..................... | .................... | .................... | .................... |
| S | 3648 hCoV-19/England/MILK-B045D5/2020\|EPI_ISL_652163\|20 | .................... | .................... | ..................... | .................... | .................... | .................... |
| S | 3649 hCoV-19/England/MILK-B04678/2020\|EPI_ISL_652164\|20 | .................... | .................... | ..................... | .................... | .................... | .................... |
| S | 3650 hCoV-19/England/MILK-B04687/2020\|EPI_ISL_652165\|20 | .................... | .................... | ..................... | .................... | ......T............. | .................... |
| S | 3651 hCoV-19/England/MILK-B046E1/2020\|EPI_ISL_652166\|20 | .................... | .................... | ..................... | .................... | .................... | .................... |
| S | 3652 hCoV-19/England/MILK-B046F0/2020\|EPI_ISL_652167\|20 | .................... | .................... | ..................... | .................... | ......T............. | .................... |
| S | 3653 hCoV-19/England/MILK-B0472A/2020\|EPI_ISL_652168\|20 | .................... | .................... | ..................... | .................... | .................... | .................... |
| S | 3654 hCoV-19/England/MILK-B04748/2020\|EPI_ISL_652169\|20 | .................... | .................... | ..................... | .................... | .................... | .................... |
| S | 3655 hCoV-19/England/MILK-B047B1/2020\|EPI_ISL_652170\|20 | .................... | .................... | ..................... | .................... | .................... | .................... |
| S | 3656 hCoV-19/England/MILK-B04872/2020\|EPI_ISL_652171\|20 | .................... | .................... | ..................... | .................... | .................... | .................... |
| S | 3657 hCoV-19/Wales/MILK-B048BE/2020\|EPI_ISL_652172\|2020 | .................... | .................... | ..................... | .................... | .................... | .................... |
| S | 3658 hCoV-19/Wales/MILK-B0497F/2020\|EPI_ISL_652173\|2020 | .................... | .................... | ..................... | .................... | .................... | .................... |
| S | 3659 hCoV-19/England/MILK-B04CA3/2020\|EPI_ISL_652174\|20 | .................... | .................... | ..................... | .................... | .................... | .................... |
| S | 3660 hCoV-19/England/MILK-B04CEF/2020\|EPI_ISL_652175\|20 | .................... | .................... | ..................... | .................... | .................... | .................... |
| S | 3661 hCoV-19/England/MILK-B04DBF/2020\|EPI_ISL_652176\|20 | .................... | .................... | ..................... | .................... | .................... | .................... |
| S | 3662 hCoV-19/England/MILK-B04DFB/2020\|EPI_ISL_652177\|20 | .................... | .................... | ..................... | .................... | .................... | .................... |
| S | 3663 hCoV-19/England/MILK-B04E43/2020\|EPI_ISL_652178\|20 | .................... | .................... | ..................... | .................... | .................... | .................... |
| S | 3664 hCoV-19/England/MILK-B04EAD/2020\|EPI_ISL_652179\|20 | .................... | .................... | ..................... | .................... | .................... | .................... |
| S | 3665 hCoV-19/USA/LA-EVTL733/2020\|EPI_ISL_653424\|2020-07 | .................... | .................... | ..................... | .................... | .................... | .................... |
| S | 3666 hCoV-19/USA/LA-EVTL734/2020\|EPI_ISL_653425\|2020-07 | .................... | .................... | ..................... | .................... | .................... | .................... |
| S | 3667 hCoV-19/USA/LA-EVTL735/2020\|EPI_ISL_653426\|2020-07 | .................... | .................... | ..................... | .................... | .................... | .................... |
| S | 3668 hCoV-19/USA/LA-EVTL736/2020\|EPI_ISL_653427\|2020-07 | .................... | .................... | ..................... | .................... | .................... | .................... |
| S | 3669 hCoV-19/USA/LA-EVTL737/2020\|EPI_ISL_653428\|2020-07 | .................... | .................... | ..................... | .................... | .................... | .................... |
| S | 3670 hCoV-19/USA/LA-EVTL738/2020\|EPI_ISL_653429\|2020-07 | .................... | .................... | ..................... | .................... | .................... | .................... |
| S | 3671 hCoV-19/USA/LA-EVTL739/2020\|EPI_ISL_653430\|2020-07 | .................... | .................... | ..................... | .................... | .................... | .................... |
| S | 3672 hCoV-19/USA/LA-EVTL740/2020\|EPI_ISL_653431\|2020-07 | .................... | .................... | ..................... | .................... | .................... | .................... |
| S | 3673 hCoV-19/USA/LA-EVTL741/2020\|EPI_ISL_653432\|2020-07 | .................... | .A.................. | ..................... | .................... | .................... | .................... |
| S | 3674 hCoV-19/USA/LA-EVTL742/2020\|EPI_ISL_653433\|2020-07 | .................... | .................... | ..................... | .................... | .................... | .................... |
| S | 3675 hCoV-19/USA/LA-EVTL743/2020\|EPI_ISL_653434\|2020-07 | .................... | .................... | ..................... | .................... | .................... | .................... |
| S | 3676 hCoV-19/USA/LA-EVTL744/2020\|EPI_ISL_653435\|2020-07 | .................... | .................... | ..................... | .................... | .................... | .................... |
| S | 3677 hCoV-19/USA/LA-EVTL745/2020\|EPI_ISL_653436\|2020-07 | .................... | .................... | ..................... | .................... | .................... | .................... |
| S | 3678 hCoV-19/USA/LA-EVTL746/2020\|EPI_ISL_653437\|2020-07 | .................... | .................... | ..................... | .................... | .................... | .................... |
| S | 3679 hCoV-19/USA/LA-EVTL747/2020\|EPI_ISL_653438\|2020-07 | .................... | .................... | ..................... | .................... | .................... | .................... |
| S | 3680 hCoV-19/USA/LA-EVTL748/2020\|EPI_ISL_653439\|2020-07 | .................... | .................... | ..................... | .................... | .................... | .................... |
| S | 3681 hCoV-19/USA/LA-EVTL750/2020\|EPI_ISL_653441\|2020-07 | .................... | .................... | ..................... | .................... | .................... | .................... |
| S | 3682 hCoV-19/USA/LA-EVTL751/2020\|EPI_ISL_653442\|2020-07 | .................... | .................... | ..................... | .................... | .................... | .................... |
| S | 3683 hCoV-19/USA/LA-EVTL752/2020\|EPI_ISL_653443\|2020-07 | .................... | .................... | ..................... | .................... | .................... | .................... |
| S | 3684 hCoV-19/USA/LA-EVTL753/2020\|EPI_ISL_653444\|2020-07 | .................... | .................... | ..................... | .................... | .................... | .................... |
| S | 3685 hCoV-19/USA/LA-EVTL754/2020\|EPI_ISL_653445\|2020-07 | .................... | .................... | ..................... | .................... | .................... | .................... |
| S | 3686 hCoV-19/USA/LA-EVTL755/2020\|EPI_ISL_653446\|2020-07 | .................... | .................... | ..................... | .................... | .................... | .................... |
| S | 3687 hCoV-19/USA/LA-EVTL756/2020\|EPI_ISL_653447\|2020-07 | .................... | .................... | ..................... | .................... | .................... | .................... |
| S | 3688 hCoV-19/USA/LA-EVTL757/2020\|EPI_ISL_653448\|2020-07 | .................... | .................... | ..................... | .................... | .................... | .................... |
| S | 3689 hCoV-19/USA/LA-EVTL758/2020\|EPI_ISL_653449\|2020-07 | .................... | .................... | ..................... | .................... | .................... | .................... |
| S | 3690 hCoV-19/USA/LA-EVTL759/2020\|EPI_ISL_653450\|2020-07 | .................... | .............A...... | ..................... | .................... | .................... | .................... |
| S | 3691 hCoV-19/USA/LA-EVTL760/2020\|EPI_ISL_653451\|2020-07 | .................... | .A.................. | ..................... | .................... | .................... | .................... |
| S | 3692 hCoV-19/USA/LA-EVTL761/2020\|EPI_ISL_653452\|2020-07 | .................... | .................... | ..................... | .................... | .................... | .................... |
| S | 3693 hCoV-19/USA/LA-EVTL762/2020\|EPI_ISL_653453\|2020-07 | .................... | .................... | ..................... | .................... | .................... | .................... |
| S | 3694 hCoV-19/USA/LA-EVTL763/2020\|EPI_ISL_653454\|2020-07 | .................... | .................... | ..................... | .................... | .................... | .................... |

| | | | | | | | |
|---|---|---|---|---|---|---|---|
| S | 3695 hCoV-19/USA/LA-EVTL764/2020\|EPI_ISL_653455\|2020-07 | ···················· | ···················· | ····················· | ···················· | ···················· | ···················· |
| S | 3696 hCoV-19/USA/LA-EVTL765/2020\|EPI_ISL_653456\|2020-07 | ···················· | ···················· | ····················· | ···················· | ···················· | ···················· |
| S | 3697 hCoV-19/USA/LA-EVTL766/2020\|EPI_ISL_653457\|2020-07 | ···················· | ···················· | ····················· | ···················· | ···················· | ···················· |
| S | 3698 hCoV-19/USA/LA-EVTL767/2020\|EPI_ISL_653458\|2020-07 | ···················· | ···················· | ····················· | ···················· | ···················· | ···················· |
| S | 3699 hCoV-19/USA/LA-EVTL768/2020\|EPI_ISL_653459\|2020-07 | ···················· | ···················· | ····················· | ···················· | ···················· | ···················· |
| S | 3700 hCoV-19/USA/LA-EVTL769/2020\|EPI_ISL_653460\|2020-07 | ···················· | ···················· | ····················· | ···················· | ···················· | ···················· |
| S | 3701 hCoV-19/USA/LA-EVTL770/2020\|EPI_ISL_653461\|2020-07 | ···················· | ···················· | ····················· | ···················· | ···················· | ···················· |
| S | 3702 hCoV-19/USA/LA-EVTL771/2020\|EPI_ISL_653462\|2020-07 | ···················· | ···················· | ····················· | ···················· | ···················· | ···················· |
| S | 3703 hCoV-19/USA/LA-EVTL772/2020\|EPI_ISL_653463\|2020-07 | ···················· | ···················· | ····················· | ···················· | ···················· | ···················· |
| S | 3704 hCoV-19/USA/LA-EVTL773/2020\|EPI_ISL_653464\|2020-07 | ···················· | ·A·················· | ····················· | ···················· | ···················· | ···················· |
| S | 3705 hCoV-19/USA/LA-EVTL775/2020\|EPI_ISL_653466\|2020-07 | ···················· | ···················· | ····················· | ···················· | ···················· | ···················· |
| S | 3706 hCoV-19/USA/LA-EVTL776/2020\|EPI_ISL_653467\|2020-07 | ···················· | ···················· | ····················· | ···················· | ···················· | ···················· |
| S | 3707 hCoV-19/USA/LA-EVTL777/2020\|EPI_ISL_653468\|2020-07 | ···················· | ···················· | ····················· | ···················· | ···················· | ···················· |
| S | 3708 hCoV-19/USA/LA-EVTL778/2020\|EPI_ISL_653469\|2020-07 | ···················· | ···················· | ····················· | ···················· | ···················· | ···················· |
| S | 3709 hCoV-19/USA/LA-EVTL779/2020\|EPI_ISL_653470\|2020-07 | ···················· | ···················· | ····················· | ···················· | ···················· | ···················· |
| S | 3710 hCoV-19/USA/LA-EVTL780/2020\|EPI_ISL_653471\|2020-07 | ···················· | ···················· | ····················· | ···················· | ···················· | ···················· |
| S | 3711 hCoV-19/USA/LA-EVTL781/2020\|EPI_ISL_653472\|2020-07 | ···················· | ···················· | ····················· | ···················· | ···················· | ···················· |
| S | 3712 hCoV-19/USA/LA-EVTL782/2020\|EPI_ISL_653473\|2020-07 | ···················· | ···················· | ····················· | ···················· | ···················· | ···················· |
| S | 3713 hCoV-19/USA/LA-EVTL783/2020\|EPI_ISL_653474\|2020-07 | ···················· | ···················· | ····················· | ···················· | ···················· | ···················· |
| S | 3714 hCoV-19/USA/LA-EVTL784/2020\|EPI_ISL_653475\|2020-07 | ···················· | ···················· | ····················· | ···················· | ···················· | ···················· |
| S | 3715 hCoV-19/USA/LA-EVTL785/2020\|EPI_ISL_653476\|2020-07 | ···················· | ···················· | ····················· | ···················· | ···················· | ···················· |
| S | 3716 hCoV-19/USA/LA-EVTL786/2020\|EPI_ISL_653477\|2020-07 | ···················· | ···················· | ····················· | ···················· | ···················· | ···················· |
| S | 3717 hCoV-19/USA/LA-EVTL787/2020\|EPI_ISL_653478\|2020-07 | ···················· | ···················· | ····················· | ···················· | ···················· | ···················· |
| S | 3718 hCoV-19/USA/LA-EVTL788/2020\|EPI_ISL_653479\|2020-07 | ···················· | ······A············· | ····················· | ···················· | ···················· | ···················· |
| S | 3719 hCoV-19/USA/LA-EVTL789/2020\|EPI_ISL_653480\|2020-07 | ···················· | ······A············· | ····················· | ···················· | ···················· | ···················· |
| S | 3720 hCoV-19/USA/LA-EVTL791/2020\|EPI_ISL_653482\|2020-07 | ···················· | ·············A······ | ····················· | ···················· | ···················· | ···················· |
| S | 3721 hCoV-19/USA/LA-EVTL792/2020\|EPI_ISL_653483\|2020-07 | ···················· | ······A············· | ····················· | ···················· | ···················· | ···················· |
| S | 3722 hCoV-19/USA/LA-EVTL793/2020\|EPI_ISL_653484\|2020-07 | ···················· | ······A············· | ····················· | ···················· | ···················· | ···················· |
| S | 3723 hCoV-19/USA/LA-EVTL794/2020\|EPI_ISL_653485\|2020-07 | ···················· | ···················· | ····················· | ···················· | ···················· | ···················· |
| S | 3724 hCoV-19/USA/LA-EVTL795/2020\|EPI_ISL_653486\|2020-07 | ···················· | ···················· | ····················· | ···················· | ···················· | ···················· |
| S | 3725 hCoV-19/USA/LA-EVTL796/2020\|EPI_ISL_653487\|2020-07 | ···················· | ···················· | ····················· | ···················· | ···················· | ···················· |
| S | 3726 hCoV-19/USA/LA-EVTL797/2020\|EPI_ISL_653488\|2020-07 | ···················· | ···················· | ····················· | ···················· | ···················· | ···················· |
| S | 3727 hCoV-19/USA/LA-EVTL798/2020\|EPI_ISL_653489\|2020-07 | ···················· | ···················· | ····················· | ···················· | ···················· | ···················· |
| S | 3728 hCoV-19/USA/LA-EVTL799/2020\|EPI_ISL_653490\|2020-07 | ···················· | ···················· | ····················· | ···················· | ···················· | ···················· |
| S | 3729 hCoV-19/USA/LA-EVTL800/2020\|EPI_ISL_653491\|2020-07 | ···················· | ···················· | ····················· | ···················· | ···················· | ···················· |
| S | 3730 hCoV-19/USA/LA-EVTL801/2020\|EPI_ISL_653492\|2020-07 | ···················· | ···················· | ····················· | ···················· | ···················· | ···················· |
| S | 3731 hCoV-19/USA/LA-EVTL802/2020\|EPI_ISL_653493\|2020-07 | ···················· | ···················· | ····················· | ···················· | ···················· | ···················· |
| S | 3732 hCoV-19/USA/LA-EVTL803/2020\|EPI_ISL_653494\|2020-07 | ···················· | ···················· | ····················· | ···················· | ···················· | ···················· |
| S | 3733 hCoV-19/USA/LA-EVTL804/2020\|EPI_ISL_653495\|2020-07 | ···················· | ···················· | ····················· | ···················· | ···················· | ···················· |
| S | 3734 hCoV-19/USA/LA-EVTL805/2020\|EPI_ISL_653496\|2020-07 | ···················· | ···················· | ····················· | ···················· | ···················· | ···················· |
| S | 3735 hCoV-19/USA/LA-EVTL806/2020\|EPI_ISL_653497\|2020-07 | ···················· | ···················· | ····················· | ···················· | ···················· | ···················· |
| S | 3736 hCoV-19/USA/LA-EVTL807/2020\|EPI_ISL_653498\|2020-07 | ···················· | ···················· | ····················· | ···················· | ···················· | ···················· |
| S | 3737 hCoV-19/USA/LA-EVTL808/2020\|EPI_ISL_653499\|2020-07 | ···················· | ···················· | ····················· | ···················· | ···················· | ···················· |
| S | 3738 hCoV-19/USA/LA-EVTL809/2020\|EPI_ISL_653500\|2020-07 | ···················· | ···················· | ····················· | ···················· | ···················· | ···················· |
| S | 3739 hCoV-19/USA/LA-EVTL810/2020\|EPI_ISL_653501\|2020-07 | ···················· | ···················· | ····················· | ···················· | ···················· | ···················· |
| S | 3740 hCoV-19/USA/LA-EVTL811/2020\|EPI_ISL_653502\|2020-07 | ···················· | ···················· | ····················· | ···················· | ···················· | ···················· |
| S | 3741 hCoV-19/USA/LA-EVTL812/2020\|EPI_ISL_653503\|2020-07 | ···················· | ···················· | ····················· | ···················· | ···················· | ···················· |
| S | 3742 hCoV-19/USA/LA-EVTL813/2020\|EPI_ISL_653504\|2020-07 | ···················· | ···················· | ····················· | ···················· | ···················· | ···················· |
| S | 3743 hCoV-19/USA/LA-EVTL814/2020\|EPI_ISL_653505\|2020-07 | ···················· | ···················· | ····················· | ···················· | ···················· | ···················· |
| S | 3744 hCoV-19/USA/LA-EVTL815/2020\|EPI_ISL_653506\|2020-07 | ···················· | ···················· | ····················· | ···················· | ···················· | ···················· |
| S | 3745 hCoV-19/USA/LA-EVTL816/2020\|EPI_ISL_653507\|2020-07 | ···················· | ···················· | ····················· | ···················· | ···················· | ···················· |
| S | 3746 hCoV-19/USA/LA-EVTL817/2020\|EPI_ISL_653508\|2020-07 | ···················· | ···················· | ····················· | ···················· | ···················· | ···················· |
| S | 3747 hCoV-19/USA/LA-EVTL818/2020\|EPI_ISL_653509\|2020-07 | ···················· | ···················· | ····················· | ···················· | ···················· | ···················· |
| S | 3748 hCoV-19/USA/LA-EVTL821/2020\|EPI_ISL_653512\|2020-07 | ···················· | ···················· | ····················· | ···················· | ···················· | ···················· |
| S | 3749 hCoV-19/USA/LA-EVTL822/2020\|EPI_ISL_653513\|2020-07 | ···················· | ···················· | ····················· | ···················· | ···················· | ···················· |
| S | 3750 hCoV-19/USA/LA-EVTL823/2020\|EPI_ISL_653514\|2020-07 | ···················· | ···················· | ····················· | ···················· | ···················· | ···················· |
| S | 3751 hCoV-19/USA/LA-EVTL824/2020\|EPI_ISL_653515\|2020-07 | ···················· | ···················· | ····················· | ···················· | ···················· | ···················· |
| S | 3752 hCoV-19/USA/LA-EVTL825/2020\|EPI_ISL_653516\|2020-07 | ···················· | ···················· | ····················· | ···················· | ···················· | ···················· |
| S | 3753 hCoV-19/USA/LA-EVTL826/2020\|EPI_ISL_653517\|2020-07 | ···················· | ···················· | ····················· | ···················· | ···················· | ···················· |
| S | 3754 hCoV-19/USA/LA-EVTL827/2020\|EPI_ISL_653518\|2020-07 | ···················· | ···················· | ····················· | ···················· | ···················· | ···················· |
| S | 3755 hCoV-19/USA/LA-EVTL828/2020\|EPI_ISL_653519\|2020-07 | ···················· | ···················· | ····················· | ···················· | ···················· | ···················· |
| S | 3756 hCoV-19/USA/LA-EVTL829/2020\|EPI_ISL_653520\|2020-07 | ···················· | ···················· | ····················· | ···················· | ···················· | ···················· |
| S | 3757 hCoV-19/USA/LA-EVTL830/2020\|EPI_ISL_653521\|2020-07 | ···················· | ···················· | ····················· | ···················· | ···················· | ···················· |
| S | 3758 hCoV-19/USA/LA-EVTL831/2020\|EPI_ISL_653522\|2020-07 | ···················· | ···················· | ····················· | ···················· | ···················· | ···················· |
| S | 3759 hCoV-19/USA/TX-DSHS-1364/2020\|EPI_ISL_654769\|2020- | ···················· | ···················· | ····················· | ···················· | ···················· | ···················· |
| S | 3760 hCoV-19/USA/TX-DSHS-1371/2020\|EPI_ISL_654770\|2020- | ···················· | ···················· | ····················· | ···················· | ···················· | ···················· |
| S | 3761 hCoV-19/USA/TX-DSHS-1380/2020\|EPI_ISL_654771\|2020- | ···················· | ···················· | ····················· | ···················· | ···················· | ···················· |
| S | 3762 hCoV-19/USA/TX-DSHS-1383/2020\|EPI_ISL_654772\|2020- | ···················· | ···················· | ····················· | ···················· | ···················· | ···················· |
| S | 3763 hCoV-19/USA/TX-DSHS-1393/2020\|EPI_ISL_654773\|2020- | ···················· | ···················· | ····················· | ···················· | ···················· | ···················· |
| S | 3764 hCoV-19/Australia/SAP548/2020\|EPI_ISL_654795\|2020- | ···················· | ·A·················· | ····················· | ···················· | ···················· | ··A················· |
| S | 3765 hCoV-19/Australia/SAP553/2020\|EPI_ISL_654799\|2020- | ···················· | ·A·················· | ····················· | ···················· | ···················· | ··A················· |
| S | 3766 hCoV-19/Australia/SAP556/2020\|EPI_ISL_654800\|2020- | ···················· | ·A·················· | ····················· | ···················· | ···················· | ··A················· |
| S | 3767 hCoV-19/Australia/SAP558/2020\|EPI_ISL_654801\|2020- | ···················· | ·A·················· | ····················· | ···················· | ···················· | ··A················· |
| S | 3768 hCoV-19/Australia/SAP559/2020\|EPI_ISL_654802\|2020- | ···················· | ·A·················· | ····················· | ···················· | ···················· | ··A················· |
| S | 3769 hCoV-19/Australia/SAP560/2020\|EPI_ISL_654803\|2020- | ···················· | ·A·················· | ····················· | ···················· | ···················· | ··A················· |
| S | 3770 hCoV-19/Australia/SAP561/2020\|EPI_ISL_654804\|2020- | ···················· | ·A·················· | ····················· | ···················· | ···················· | ··A················· |
| S | 3771 hCoV-19/Australia/SAP562/2020\|EPI_ISL_654805\|2020- | ···················· | ·A·················· | ····················· | ···················· | ···················· | ··A················· |
| S | 3772 hCoV-19/Australia/SAP563/2020\|EPI_ISL_654806\|2020- | ···················· | ·A·················· | ····················· | ···················· | ···················· | ··A················· |
| S | 3773 hCoV-19/Australia/SAP564/2020\|EPI_ISL_654807\|2020- | ···················· | ·A·················· | ····················· | ···················· | ···················· | ··A················· |
| S | 3774 hCoV-19/Australia/SAP565/2020\|EPI_ISL_654808\|2020- | ···················· | ·A·················· | ····················· | ···················· | ···················· | ··A················· |

| | | | | | | | |
|---|---|---|---|---|---|---|---|
| S | 3775 hCoV-19/Australia/SAP566/2020\|EPI_ISL_654809\|2020- | ···················· | ·A·················· | ····················· | ···················· | ···················· | ···A················ |
| S | 3776 hCoV-19/Singapore/1412/2020\|EPI_ISL_654810\|2020-11 | ···················· | ···················· | ····················· | ···················· | ···················· | ···················· |
| S | 3777 hCoV-19/Singapore/1413/2020\|EPI_ISL_654811\|2020-11 | ···················· | ·A·················· | ····················· | ···················· | ···················· | ···A················ |
| S | 3778 hCoV-19/Singapore/1414/2020\|EPI_ISL_654812\|2020-11 | ···················· | ·A·················· | ····················· | ···················· | ···················· | ···A················ |
| S | 3779 hCoV-19/Singapore/1415/2020\|EPI_ISL_654813\|2020-11 | ···················· | ···················· | ····················· | ···················· | ···················· | ···················· |
| S | 3780 hCoV-19/Singapore/1416/2020\|EPI_ISL_654814\|2020-11 | ···················· | ·A·················· | ····················· | ···················· | ···················· | ···A················ |
| S | 3781 hCoV-19/Singapore/1417/2020\|EPI_ISL_654815\|2020-11 | ···················· | ·A·················· | ····················· | ···················· | ···················· | ···A················ |
| S | 3782 hCoV-19/Singapore/1414_2/2020\|EPI_ISL_654816\|2020- | ···················· | ·A·················· | ····················· | ···················· | ···················· | ···A················ |
| S | 3783 hCoV-19/Singapore/1418/2020\|EPI_ISL_654817\|2020-11 | ···················· | ···················· | ····················· | ···················· | ···················· | ···················· |
| S | 3784 hCoV-19/Ukraine/Chernivtsi_860/2020\|EPI_ISL_654818 | ···················· | ···················· | ····················· | ···················· | ···················· | ···················· |
| S | 3785 hCoV-19/Ukraine/Chernivtsi_861/2020\|EPI_ISL_654819 | ···················· | ···················· | ····················· | ···················· | ···················· | ···················· |
| S | 3786 hCoV-19/Ukraine/Lutsk_824/2020\|EPI_ISL_654820\|2020 | ···················· | ···················· | ····················· | ···················· | ···················· | ···················· |
| S | 3787 hCoV-19/England/QEUH-B4086B/2020\|EPI_ISL_657070\|20 | ···················· | ···················· | ····················· | ···················· | ···················· | ···················· |
| S | 3788 hCoV-19/England/QEUH-B40519/2020\|EPI_ISL_657072\|20 | ···················· | ···················· | ····················· | ···················· | ···················· | ···················· |
| S | 3789 hCoV-19/England/QEUH-B42140/2020\|EPI_ISL_657073\|20 | ···················· | ···················· | ····················· | ···················· | ···················· | ···················· |
| S | 3790 hCoV-19/England/QEUH-B425F9/2020\|EPI_ISL_657074\|20 | ···················· | ···················· | ····················· | ···················· | ···················· | ···················· |
| S | 3791 hCoV-19/England/QEUH-B3FE12/2020\|EPI_ISL_657075\|20 | ···················· | ·A·················· | ····················· | ···················· | ···················· | ···A················ |
| S | 3792 hCoV-19/England/QEUH-B43710/2020\|EPI_ISL_657076\|20 | ···················· | ···················· | ····················· | ···················· | ···················· | ···················· |
| S | 3793 hCoV-19/England/QEUH-B42474/2020\|EPI_ISL_657077\|20 | ···················· | ···················· | ····················· | ···················· | ···················· | ···················· |
| S | 3794 hCoV-19/England/QEUH-B437B6/2020\|EPI_ISL_657078\|20 | ···················· | ···················· | ····················· | ···················· | ···················· | ···················· |
| S | 3795 hCoV-19/England/QEUH-B421B9/2020\|EPI_ISL_657079\|20 | ···················· | ·A·················· | ····················· | ···················· | ···················· | ···A················ |
| S | 3796 hCoV-19/England/QEUH-B4375C/2020\|EPI_ISL_657080\|20 | ···················· | ···················· | ····················· | ···················· | ···················· | ···················· |
| S | 3797 hCoV-19/England/QEUH-B4373E/2020\|EPI_ISL_657081\|20 | ···················· | ···················· | ····················· | ···················· | ···················· | ···················· |
| S | 3798 hCoV-19/England/QEUH-B3FEE5/2020\|EPI_ISL_657083\|20 | ···················· | ···················· | ····················· | ···················· | ···················· | ···················· |
| S | 3799 hCoV-19/England/QEUH-B436D7/2020\|EPI_ISL_657086\|20 | ···················· | ···················· | ····················· | ···················· | ···················· | ···················· |
| S | 3800 hCoV-19/England/QEUH-B438D1/2020\|EPI_ISL_657087\|20 | ···················· | ···················· | ····················· | ···················· | ···················· | ···················· |
| S | 3801 hCoV-19/England/QEUH-B3FE03/2020\|EPI_ISL_657088\|20 | ···················· | ···················· | ····················· | ···················· | ···················· | ···················· |
| S | 3802 hCoV-19/England/QEUH-B4215F/2020\|EPI_ISL_657089\|20 | ···················· | ···················· | ····················· | ···················· | ······T············· | ···················· |
| S | 3803 hCoV-19/England/QEUH-B426AB/2020\|EPI_ISL_657090\|20 | ···················· | ···················· | ····················· | ···················· | ···················· | ···················· |
| S | 3804 hCoV-19/England/QEUH-B423B3/2020\|EPI_ISL_657091\|20 | ···················· | ·A·················· | ····················· | ···················· | ···················· | ···················· |
| S | 3805 hCoV-19/England/QEUH-B424B0/2020\|EPI_ISL_657092\|20 | ···················· | ···················· | ····················· | ···················· | ···················· | ···················· |
| S | 3806 hCoV-19/England/QEUH-B4234A/2020\|EPI_ISL_657094\|20 | ···················· | ···················· | ····················· | ···················· | ···················· | ···················· |
| S | 3807 hCoV-19/England/QEUH-B4218C/2020\|EPI_ISL_657095\|20 | ···················· | ···················· | ····················· | ···················· | ···················· | ···················· |
| S | 3808 hCoV-19/England/QEUH-B3FFD3/2020\|EPI_ISL_657096\|20 | ···················· | ···················· | ····················· | ···················· | ···················· | ···················· |
| S | 3809 hCoV-19/England/QEUH-B4026D/2020\|EPI_ISL_657097\|20 | ···················· | ···················· | ····················· | ···················· | ···················· | ···················· |
| S | 3810 hCoV-19/England/QEUH-B3FFC4/2020\|EPI_ISL_657098\|20 | ···················· | ···················· | ····················· | ···················· | ···················· | ···················· |
| S | 3811 hCoV-19/England/QEUH-B40054/2020\|EPI_ISL_657099\|20 | ···················· | ···················· | ····················· | ···················· | ···················· | ···················· |
| S | 3812 hCoV-19/England/MILK-B5A6FB/2020\|EPI_ISL_658300\|20 | ···················· | ···················· | ····················· | ···················· | ···················· | ···················· |
| S | 3813 hCoV-19/England/MILK-B55BB5/2020\|EPI_ISL_658301\|20 | ···················· | ···················· | ····················· | ···················· | ···················· | ···················· |
| S | 3814 hCoV-19/England/MILK-B5A497/2020\|EPI_ISL_658302\|20 | ···················· | ···················· | ····················· | ···················· | ···················· | ···················· |
| S | 3815 hCoV-19/England/CAMC-B51BE6/2020\|EPI_ISL_658304\|20 | ···················· | ···················· | ····················· | ···················· | ···················· | ···················· |
| S | 3816 hCoV-19/England/CAMC-B51973/2020\|EPI_ISL_658305\|20 | ···················· | ·A·················· | ····················· | ···················· | ···················· | ···A················ |
| S | 3817 hCoV-19/England/MILK-B55E16/2020\|EPI_ISL_658306\|20 | ···················· | ···················· | ····················· | ···················· | ···················· | ···················· |
| S | 3818 hCoV-19/England/CAMC-B54837/2020\|EPI_ISL_658310\|20 | ···················· | ···················· | ····················· | ···················· | ···················· | ···················· |
| S | 3819 hCoV-19/England/CAMC-B51AE9/2020\|EPI_ISL_658311\|20 | ···················· | ···················· | ····················· | ···················· | ···················· | ···················· |
| S | 3820 hCoV-19/England/CAMC-B5480A/2020\|EPI_ISL_658313\|20 | ···················· | ···················· | ····················· | ···················· | ···················· | ···················· |
| S | 3821 hCoV-19/England/MILK-B55C3A/2020\|EPI_ISL_658315\|20 | ···················· | ·A·················· | ····················· | ···················· | ···················· | ···A················ |
| S | 3822 hCoV-19/England/CAMC-B5463D/2020\|EPI_ISL_658316\|20 | ···················· | ···················· | ····················· | ···················· | ···················· | ···················· |
| S | 3823 hCoV-19/England/CAMC-B519DD/2020\|EPI_ISL_658317\|20 | ···················· | ···················· | ····················· | ···················· | ···················· | ···················· |
| S | 3824 hCoV-19/England/MILK-B55CEF/2020\|EPI_ISL_658318\|20 | ···················· | ···················· | ····················· | ···················· | ···················· | ···················· |
| S | 3825 hCoV-19/England/CAMC-B32C7F/2020\|EPI_ISL_659629\|20 | ···················· | ···················· | ····················· | ···················· | ···················· | ···················· |
| S | 3826 hCoV-19/England/CAMC-B32DF4/2020\|EPI_ISL_659631\|20 | ···················· | ···················· | ····················· | ···················· | ···················· | ···················· |
| S | 3827 hCoV-19/England/CAMC-B3478B/2020\|EPI_ISL_659632\|20 | ···················· | ···················· | ····················· | ···················· | ······T············· | ···················· |
| S | 3828 hCoV-19/England/CAMC-B344DF/2020\|EPI_ISL_659634\|20 | ···················· | ···················· | ····················· | ···················· | ···················· | ···················· |
| S | 3829 hCoV-19/England/CAMC-B344C0/2020\|EPI_ISL_659635\|20 | ···················· | ·A·················· | ····················· | ···················· | ···················· | ···A················ |
| S | 3830 hCoV-19/England/CAMC-B32A84/2020\|EPI_ISL_659640\|20 | ···················· | ···················· | ····················· | ···················· | ······T············· | ···················· |
| S | 3831 hCoV-19/England/CAMC-B3493A/2020\|EPI_ISL_659641\|20 | ···················· | ···················· | ····················· | ···················· | ···················· | ···················· |
| S | 3832 hCoV-19/England/CAMC-B3567E/2020\|EPI_ISL_659643\|20 | ···················· | ···················· | ····················· | ···················· | ···················· | ···················· |
| S | 3833 hCoV-19/England/CAMC-B34475/2020\|EPI_ISL_659644\|20 | ···················· | ···················· | ····················· | ···················· | ···················· | ···················· |
| S | 3834 hCoV-19/England/CAMC-B34BF8/2020\|EPI_ISL_659646\|20 | ···················· | ···················· | ····················· | ···················· | ···················· | ···················· |
| S | 3835 hCoV-19/England/CAMC-B35535/2020\|EPI_ISL_659647\|20 | ···················· | ·A·················· | ····················· | ···················· | ···················· | ···A················ |
| S | 3836 hCoV-19/England/CAMC-B3574E/2020\|EPI_ISL_659648\|20 | ···················· | ···················· | ····················· | ···················· | ···················· | ···················· |
| S | 3837 hCoV-19/England/CAMC-B32CAC/2020\|EPI_ISL_659649\|20 | ···················· | ···················· | ····················· | ···················· | ···················· | ···················· |
| S | 3838 hCoV-19/South Africa/KRISP-EC-K004578/2020\|EPI_ISL | ···················· | ···················· | ····················· | ···················· | ···················· | ···················· |
| S | 3839 hCoV-19/South Africa/KRISP-K004588/2020\|EPI_ISL_66 | ···················· | ···················· | ····················· | ···················· | ···················· | ···················· |
| S | 3840 hCoV-19/South Africa/KRISP-K004589/2020\|EPI_ISL_66 | ···················· | ···················· | ····················· | ···················· | ···················· | ···················· |
| S | 3841 hCoV-19/Wales/MILK-B05370/2020\|EPI_ISL_652180\|2020 | ···················· | ···················· | ····················· | ···················· | ···················· | ···················· |
| S | 3842 hCoV-19/England/MILK-B0546E/2020\|EPI_ISL_652181\|20 | ···················· | ···················· | ····················· | ···················· | ···················· | ···················· |
| S | 3843 hCoV-19/England/MILK-B054D7/2020\|EPI_ISL_652182\|20 | ···················· | ···················· | ····················· | ···················· | ···················· | ···················· |
| S | 3844 hCoV-19/England/MILK-B054E6/2020\|EPI_ISL_652183\|20 | ···················· | ···················· | ····················· | ···················· | ···················· | ···················· |
| S | 3845 hCoV-19/England/MILK-B0556B/2020\|EPI_ISL_652184\|20 | ···················· | ···················· | ····················· | ···················· | ···················· | ···················· |
| S | 3846 hCoV-19/England/MILK-B05589/2020\|EPI_ISL_652185\|20 | ···················· | ···················· | ····················· | ···················· | ···················· | ···················· |
| S | 3847 hCoV-19/Wales/MILK-B0563B/2020\|EPI_ISL_652186\|2020 | ···················· | ···················· | ····················· | ···················· | ···················· | ···················· |
| S | 3848 hCoV-19/England/MILK-B05659/2020\|EPI_ISL_652187\|20 | ···················· | ···················· | ····················· | ···················· | ······T············· | ···················· |
| S | 3849 hCoV-19/England/MILK-B05695/2020\|EPI_ISL_652188\|20 | ···················· | ···················· | ····················· | ···················· | ···················· | ···················· |
| S | 3850 hCoV-19/England/MILK-B056B3/2020\|EPI_ISL_652189\|20 | ···················· | ···················· | ····················· | ···················· | ···················· | ···················· |
| S | 3851 hCoV-19/Wales/MILK-B056C2/2020\|EPI_ISL_652190\|2020 | ···················· | ···················· | ····················· | ···················· | ···················· | ···················· |
| S | 3852 hCoV-19/England/MILK-B056FF/2020\|EPI_ISL_652191\|20 | ···················· | ···················· | ····················· | ···················· | ···················· | ···················· |
| S | 3853 hCoV-19/England/MILK-B0570B/2020\|EPI_ISL_652192\|20 | ···················· | ···················· | ····················· | ···················· | ···················· | ···················· |
| S | 3854 hCoV-19/England/MILK-B05738/2020\|EPI_ISL_652193\|20 | ···················· | ···················· | ····················· | ···················· | ···················· | ···················· |

| | | | | | | | | |
|---|---|---|---|---|---|---|---|---|
| S | 3855 hCoV-19/England/MILK-B05747/2020\|EPI_ISL_652194\|20 | ···················· | ···················· | ····················· | ···················· | ···················· | ···················· | |
| S | 3856 hCoV-19/England/MILK-B05756/2020\|EPI_ISL_652195\|20 | ···················· | ···················· | ····················· | ···················· | ···················· | ···················· | |
| S | 3857 hCoV-19/England/MILK-B05765/2020\|EPI_ISL_652196\|20 | ···················· | ···················· | ····················· | ···················· | ···················· | ···················· | |
| S | 3858 hCoV-19/England/MILK-B05783/2020\|EPI_ISL_652197\|20 | ···················· | ···················· | ····················· | ···················· | ···················· | ···················· | |
| S | 3859 hCoV-19/England/MILK-B05808/2020\|EPI_ISL_652198\|20 | ···················· | ···················· | ····················· | ···················· | ···················· | ···················· | |
| S | 3860 hCoV-19/England/MILK-B05817/2020\|EPI_ISL_652199\|20 | ···················· | ···················· | ····················· | ···················· | ···················· | ···················· | |
| S | 3861 hCoV-19/USA/LA-EVTL710/2020\|EPI_ISL_653402\|2020-07 | ···················· | ···················· | ····················· | ···················· | ···················· | ···················· | |
| S | 3862 hCoV-19/USA/LA-EVTL711/2020\|EPI_ISL_653403\|2020-07 | ···················· | ···················· | ····················· | ···················· | ···················· | ···················· | |
| S | 3863 hCoV-19/USA/LA-EVTL712/2020\|EPI_ISL_653404\|2020-07 | ···················· | ···················· | ····················· | ···················· | ···················· | ···················· | |
| S | 3864 hCoV-19/USA/LA-EVTL713/2020\|EPI_ISL_653405\|2020-07 | ···················· | ···················· | ····················· | ···················· | ···················· | ···················· | |
| S | 3865 hCoV-19/USA/LA-EVTL714/2020\|EPI_ISL_653406\|2020-07 | ···················· | ·A·················· | ····················· | ···················· | ···················· | ···················· | |
| S | 3866 hCoV-19/USA/LA-EVTL715/2020\|EPI_ISL_653407\|2020-07 | ···················· | ······A············· | ····················· | ···················· | ···················· | ···················· | |
| S | 3867 hCoV-19/USA/LA-EVTL716/2020\|EPI_ISL_653408\|2020-07 | ···················· | ···················· | ····················· | ···················· | ···················· | ···················· | |
| S | 3868 hCoV-19/USA/LA-EVTL717/2020\|EPI_ISL_653409\|2020-07 | ···················· | ···················· | ····················· | ···················· | ···················· | ···················· | |
| S | 3869 hCoV-19/USA/LA-EVTL718/2020\|EPI_ISL_653410\|2020-07 | ···················· | ···················· | ····················· | ···················· | ···················· | ···················· | |
| S | 3870 hCoV-19/USA/LA-EVTL719/2020\|EPI_ISL_653411\|2020-07 | ···················· | ···················· | ····················· | ···················· | ···················· | ···················· | |
| S | 3871 hCoV-19/USA/LA-EVTL720/2020\|EPI_ISL_653412\|2020-07 | ···················· | ···················· | ····················· | ···················· | ···················· | ···················· | |
| S | 3872 hCoV-19/USA/LA-EVTL721/2020\|EPI_ISL_653413\|2020-07 | ···················· | ···················· | ····················· | ···················· | ···················· | ···················· | |
| S | 3873 hCoV-19/USA/LA-EVTL722/2020\|EPI_ISL_653414\|2020-07 | ···················· | ·A·················· | ····················· | ···················· | ···················· | ···················· | |
| S | 3874 hCoV-19/USA/LA-EVTL723/2020\|EPI_ISL_653415\|2020-07 | ···················· | ···················· | ····················· | ···················· | ···················· | ···················· | |
| S | 3875 hCoV-19/USA/LA-EVTL724/2020\|EPI_ISL_653416\|2020-07 | ···················· | ···················· | ····················· | ···················· | ···················· | ···················· | |
| S | 3876 hCoV-19/USA/LA-EVTL725/2020\|EPI_ISL_653417\|2020-07 | ···················· | ·A·················· | ····················· | ···················· | ···················· | ···················· | |
| S | 3877 hCoV-19/USA/LA-EVTL726/2020\|EPI_ISL_653418\|2020-07 | ···················· | ···················· | ····················· | ···················· | ···················· | ···················· | |
| S | 3878 hCoV-19/USA/LA-EVTL727/2020\|EPI_ISL_653419\|2020-07 | ···················· | ···················· | ····················· | ···················· | ···················· | ···················· | |
| S | 3879 hCoV-19/USA/LA-EVTL729/2020\|EPI_ISL_653420\|2020-07 | ···················· | ···················· | ····················· | ···················· | ···················· | ···················· | |
| S | 3880 hCoV-19/USA/LA-EVTL730/2020\|EPI_ISL_653421\|2020-07 | ···················· | ···················· | ····················· | ···················· | ···················· | ···················· | |
| S | 3881 hCoV-19/USA/LA-EVTL731/2020\|EPI_ISL_653422\|2020-07 | ···················· | ···················· | ····················· | ···················· | ···················· | ···················· | |
| S | 3882 hCoV-19/USA/LA-EVTL732/2020\|EPI_ISL_653423\|2020-07 | ···················· | ···················· | ····················· | ···················· | ···················· | ···················· | |
| S | 3883 hCoV-19/USA/MN-MDH-2030/2020\|EPI_ISL_654711\|2020-1 | ···················· | ···················· | ····················· | ···················· | ···················· | ···················· | |
| S | 3884 hCoV-19/USA/MN-MDH-2031/2020\|EPI_ISL_654712\|2020-1 | ···················· | ···················· | ····················· | ···················· | NNN················· | ···················· | 1 |
| S | 3885 hCoV-19/USA/MN-MDH-2032/2020\|EPI_ISL_654713\|2020-1 | ···················· | ···················· | ····················· | ···················· | ···················· | ···················· | |
| S | 3886 hCoV-19/USA/MN-MDH-2033/2020\|EPI_ISL_654714\|2020-1 | ···················· | ···················· | ····················· | ···················· | ···················· | ···················· | |
| S | 3887 hCoV-19/USA/MN-MDH-2036/2020\|EPI_ISL_654717\|2020-1 | ···················· | ···················· | ····················· | ···················· | ···················· | ···················· | |
| S | 3888 hCoV-19/USA/MN-MDH-2037/2020\|EPI_ISL_654718\|2020-1 | ···················· | ···················· | ····················· | ···················· | ···················· | ···················· | |
| S | 3889 hCoV-19/USA/MN-MDH-2039/2020\|EPI_ISL_654720\|2020-1 | ···················· | ···················· | ····················· | ···················· | ···················· | ···················· | |
| S | 3890 hCoV-19/USA/MN-MDH-2040/2020\|EPI_ISL_654721\|2020-1 | ···················· | ···················· | ····················· | ···················· | ···················· | ···················· | |
| S | 3891 hCoV-19/England/CAMC-B548EC/2020\|EPI_ISL_658202\|20 | ···················· | ···················· | ····················· | ···················· | ···················· | ···················· | |
| S | 3892 hCoV-19/England/CAMC-B5499E/2020\|EPI_ISL_658203\|20 | ···················· | ···················· | ····················· | ···················· | ···················· | ···················· | |
| S | 3893 hCoV-19/England/MILK-B5A41F/2020\|EPI_ISL_658204\|20 | ···················· | ···················· | ····················· | ···················· | ···················· | ···················· | |
| S | 3894 hCoV-19/England/CAMC-B549E9/2020\|EPI_ISL_658206\|20 | ···················· | ···················· | ····················· | ···················· | ···················· | ···················· | |
| S | 3895 hCoV-19/England/CAMC-B54970/2020\|EPI_ISL_658207\|20 | ···················· | ···················· | ····················· | ···················· | ···················· | ···················· | |
| S | 3896 hCoV-19/England/CAMC-B516F4/2020\|EPI_ISL_658209\|20 | ···················· | ···················· | ····················· | ···················· | ···················· | ···················· | |
| S | 3897 hCoV-19/England/MILK-B5A2CA/2020\|EPI_ISL_658210\|20 | ···················· | ···················· | ····················· | ···················· | ···················· | ···················· | |
| S | 3898 hCoV-19/England/MILK-B5A4E2/2020\|EPI_ISL_658211\|20 | ···················· | ···················· | ····················· | ···················· | ······T············· | ···················· | |
| S | 3899 hCoV-19/England/CAMC-B5461F/2020\|EPI_ISL_658212\|20 | ···················· | ···················· | ····················· | ···················· | ···················· | ···················· | |
| S | 3900 hCoV-19/England/MILK-B5A2D9/2020\|EPI_ISL_658213\|20 | ···················· | ···················· | ····················· | ···················· | ···················· | ···················· | |
| S | 3901 hCoV-19/England/MILK-B55FC8/2020\|EPI_ISL_658216\|20 | ···················· | ···················· | ····················· | ···················· | ···················· | ···················· | |
| S | 3902 hCoV-19/England/CAMC-B5464C/2020\|EPI_ISL_658218\|20 | ···················· | ···················· | ····················· | ···················· | ···················· | ···················· | |
| S | 3903 hCoV-19/England/MILK-B55BC4/2020\|EPI_ISL_658219\|20 | ···················· | ···················· | ····················· | ···················· | ···················· | ···················· | |
| S | 3904 hCoV-19/England/CAMC-B51B31/2020\|EPI_ISL_658220\|20 | ···················· | ···················· | ····················· | ···················· | ···················· | ···················· | |
| S | 3905 hCoV-19/England/CAMC-B51876/2020\|EPI_ISL_658221\|20 | ···················· | ···················· | ····················· | ···················· | ···················· | ···················· | |
| S | 3906 hCoV-19/England/MILK-B5A34F/2020\|EPI_ISL_658222\|20 | ···················· | ···················· | ····················· | ···················· | ···················· | ···················· | |
| S | 3907 hCoV-19/England/MILK-B5A831/2020\|EPI_ISL_658223\|20 | ···················· | ···················· | ····················· | ···················· | ···················· | ···················· | |
| S | 3908 hCoV-19/England/CAMC-B51BD7/2020\|EPI_ISL_658224\|20 | ···················· | ···················· | ····················· | ···················· | ···················· | ···················· | |
| S | 3909 hCoV-19/England/MILK-B55F6E/2020\|EPI_ISL_658225\|20 | ···················· | ···················· | ····················· | ···················· | ···················· | ···················· | |
| S | 3910 hCoV-19/England/CAMC-B51946/2020\|EPI_ISL_658227\|20 | ···················· | ···················· | ····················· | ···················· | ······T············· | ···················· | |
| S | 3911 hCoV-19/England/CAMC-B549CB/2020\|EPI_ISL_658228\|20 | ···················· | ···················· | ····················· | ···················· | ···················· | ···················· | |
| S | 3912 hCoV-19/England/MILK-B5A7CB/2020\|EPI_ISL_658229\|20 | ···················· | ···················· | ····················· | ···················· | ···················· | ···················· | |
| S | 3913 hCoV-19/England/MILK-B5605B/2020\|EPI_ISL_658230\|20 | ···················· | ···················· | ····················· | ···················· | ···················· | ···················· | |
| S | 3914 hCoV-19/England/CAMC-B547A3/2020\|EPI_ISL_658231\|20 | ···················· | ···················· | ····················· | ···················· | ···················· | ···················· | |
| S | 3915 hCoV-19/England/MILK-B5A5C1/2020\|EPI_ISL_658233\|20 | ···················· | ···················· | ····················· | ···················· | ···················· | ···················· | |
| S | 3916 hCoV-19/England/CAMC-B51AAD/2020\|EPI_ISL_658234\|20 | ···················· | ···················· | ····················· | ···················· | ···················· | ···················· | |
| S | 3917 hCoV-19/England/MILK-B5A2AC/2020\|EPI_ISL_658235\|20 | ···················· | ···················· | ····················· | ···················· | ···················· | ···················· | |
| S | 3918 hCoV-19/England/CAMC-B51919/2020\|EPI_ISL_658236\|20 | ···················· | ···················· | ····················· | ···················· | ···················· | ···················· | |
| S | 3919 hCoV-19/England/MILK-B560C4/2020\|EPI_ISL_658237\|20 | ···················· | ···················· | ····················· | ···················· | ···················· | ···················· | |
| S | 3920 hCoV-19/England/MILK-B5A7F8/2020\|EPI_ISL_658238\|20 | ···················· | ···················· | ····················· | ···················· | ···················· | ···················· | |
| S | 3921 hCoV-19/England/CAMC-B5190A/2020\|EPI_ISL_658239\|20 | ···················· | ···················· | ····················· | ···················· | ···················· | ···················· | |
| S | 3922 hCoV-19/England/MILK-B5A50D/2020\|EPI_ISL_658241\|20 | ···················· | ···················· | ····················· | ···················· | ···················· | ···················· | |
| S | 3923 hCoV-19/England/CAMC-B5165E/2020\|EPI_ISL_658242\|20 | ···················· | ···················· | ····················· | ···················· | ······T············· | ···················· | |
| S | 3924 hCoV-19/England/MILK-B5A594/2020\|EPI_ISL_658243\|20 | ···················· | ···················· | ····················· | ···················· | ···················· | ···················· | |
| S | 3925 hCoV-19/England/MILK-B5A761/2020\|EPI_ISL_658244\|20 | ···················· | ···················· | ····················· | ···················· | ···················· | ···················· | |
| S | 3926 hCoV-19/England/MILK-B5A39A/2020\|EPI_ISL_658245\|20 | ···················· | ···················· | ····················· | ···················· | ···················· | ···················· | |
| S | 3927 hCoV-19/England/CAMC-B5498F/2020\|EPI_ISL_658246\|20 | ···················· | ·A·················· | ····················· | ···················· | ···················· | ···A················ | |
| S | 3928 hCoV-19/England/MILK-B5A3E5/2020\|EPI_ISL_658247\|20 | ···················· | ···················· | ····················· | ···················· | ···················· | ···················· | |
| S | 3929 hCoV-19/England/MILK-B5A79E/2020\|EPI_ISL_658248\|20 | ···················· | ···················· | ····················· | ···················· | ···················· | ···················· | |
| S | 3930 hCoV-19/England/MILK-B55C94/2020\|EPI_ISL_658251\|20 | ···················· | ···················· | ····················· | ···················· | ···················· | ···················· | |
| S | 3931 hCoV-19/England/CAMC-B51885/2020\|EPI_ISL_658252\|20 | ···················· | ···················· | ····················· | ···················· | ···················· | ···················· | |
| S | 3932 hCoV-19/England/MILK-B5A725/2020\|EPI_ISL_658254\|20 | ···················· | ···················· | ····················· | ···················· | ···················· | ···················· | |
| S | 3933 hCoV-19/England/MILK-B5A4D3/2020\|EPI_ISL_658255\|20 | ···················· | ···················· | ····················· | ···················· | ···················· | ···················· | |
| S | 3934 hCoV-19/England/CAMC-B54846/2020\|EPI_ISL_658256\|20 | ···················· | ···················· | ····················· | ···················· | ···················· | ···················· | |

| | | | | | | | |
|---|---|---|---|---|---|---|---|
| S | 3935 hCoV-19/England/MILK-B5A242/2020\|EPI_ISL_658257\|20 | ···················· | ···················· | ···················· | ···················· | ···················· | ···················· |
| S | 3936 hCoV-19/England/CAMC-B54934/2020\|EPI_ISL_658258\|20 | ···················· | ···················· | ···················· | ···················· | ···················· | ···················· |
| S | 3937 hCoV-19/England/MILK-B5A37C/2020\|EPI_ISL_658261\|20 | ···················· | ···················· | ···················· | ···················· | ···················· | ···················· |
| S | 3938 hCoV-19/England/MILK-B5602E/2020\|EPI_ISL_658262\|20 | ···················· | ···················· | ···················· | ···················· | ···················· | ···················· |
| S | 3939 hCoV-19/England/CAMC-B549BC/2020\|EPI_ISL_658263\|20 | ···················· | ···················· | ···················· | ···················· | ···················· | ···················· |
| S | 3940 hCoV-19/England/MILK-B5A29D/2020\|EPI_ISL_658264\|20 | ···················· | ···················· | ···················· | ···················· | ···················· | ···················· |
| S | 3941 hCoV-19/England/MILK-B55C85/2020\|EPI_ISL_658265\|20 | ···················· | ···················· | ···················· | ···················· | ···················· | ···················· |
| S | 3942 hCoV-19/England/CAMC-B51849/2020\|EPI_ISL_658266\|20 | ···················· | ···················· | ···················· | ···················· | ···················· | ···················· |
| S | 3943 hCoV-19/England/MILK-B55F7D/2020\|EPI_ISL_658268\|20 | ···················· | ···················· | ···················· | ···················· | ···················· | ···················· |
| S | 3944 hCoV-19/England/CAMC-B54952/2020\|EPI_ISL_658270\|20 | ···················· | ···················· | ···················· | ···················· | ···················· | ···················· |
| S | 3945 hCoV-19/England/CAMC-B54864/2020\|EPI_ISL_658271\|20 | ···················· | ···················· | ···················· | ···················· | ······T············· | ···················· |
| S | 3946 hCoV-19/England/CAMC-B51991/2020\|EPI_ISL_658272\|20 | ···················· | ·A·················· | ···················· | ···················· | ···················· | ··A················· |
| S | 3947 hCoV-19/England/MILK-B55BE2/2020\|EPI_ISL_658273\|20 | ···················· | ···················· | ···················· | ···················· | ···················· | ···················· |
| S | 3948 hCoV-19/England/CAMC-B51B04/2020\|EPI_ISL_658274\|20 | ···················· | ···················· | ···················· | ···················· | ···················· | ···················· |
| S | 3949 hCoV-19/England/CAMC-B51A9E/2020\|EPI_ISL_658275\|20 | ···················· | ···················· | ···················· | ···················· | ···················· | ···················· |
| S | 3950 hCoV-19/England/CAMC-B51A70/2020\|EPI_ISL_658276\|20 | ···················· | ···················· | ···················· | ···················· | ···················· | ···················· |
| S | 3951 hCoV-19/England/MILK-B5A60A/2020\|EPI_ISL_658277\|20 | ···················· | ···················· | ···················· | ···················· | ···················· | ···················· |
| S | 3952 hCoV-19/England/CAMC-B5465B/2020\|EPI_ISL_658278\|20 | ···················· | ·A·················· | ···················· | ···················· | ···················· | ···················· |
| S | 3953 hCoV-19/England/CAMC-B546B5/2020\|EPI_ISL_658279\|20 | ···················· | ···················· | ···················· | ···················· | ···················· | ···················· |
| S | 3954 hCoV-19/England/MILK-B5A3F4/2020\|EPI_ISL_658308\|20 | ···················· | ···················· | ···················· | ···················· | ···················· | ···················· |
| S | 3955 hCoV-19/England/MILK-B5A734/2020\|EPI_ISL_658309\|20 | ···················· | ···················· | ···················· | ···················· | ···················· | ···················· |
| S | 3956 hCoV-19/England/CAMC-B3291E/2020\|EPI_ISL_659529\|20 | ···················· | ···················· | ···················· | ···················· | ···················· | ···················· |
| S | 3957 hCoV-19/England/CAMC-B34509/2020\|EPI_ISL_659531\|20 | ···················· | ···················· | ···················· | ···················· | ···················· | ···················· |
| S | 3958 hCoV-19/England/CAMC-B346AC/2020\|EPI_ISL_659532\|20 | ···················· | ···················· | ···················· | ···················· | ···················· | ···················· |
| S | 3959 hCoV-19/England/CAMC-B347D6/2020\|EPI_ISL_659533\|20 | ···················· | ···················· | ···················· | ···················· | ···················· | ···················· |
| S | 3960 hCoV-19/England/CAMC-B34581/2020\|EPI_ISL_659534\|20 | ···················· | ···················· | ···················· | ···················· | ······T············· | ···················· |
| S | 3961 hCoV-19/England/CAMC-B34B52/2020\|EPI_ISL_659535\|20 | ···················· | ···················· | ···················· | ···················· | ···················· | ···················· |
| S | 3962 hCoV-19/England/CAMC-B35720/2020\|EPI_ISL_659536\|20 | ···················· | ···················· | ···················· | ···················· | ···················· | ···················· |
| S | 3963 hCoV-19/England/CAMC-B32D5E/2020\|EPI_ISL_659537\|20 | ···················· | ···················· | ···················· | ···················· | ···················· | ···················· |
| S | 3964 hCoV-19/England/CAMC-B343B4/2020\|EPI_ISL_659538\|20 | ···················· | ···················· | ···················· | ···················· | ···················· | ···················· |
| S | 3965 hCoV-19/England/CAMC-B35702/2020\|EPI_ISL_659539\|20 | ···················· | ···················· | ···················· | ···················· | ···················· | ···················· |
| S | 3966 hCoV-19/England/CAMC-B35641/2020\|EPI_ISL_659540\|20 | ···················· | ···················· | ···················· | ···················· | ···················· | ···················· |
| S | 3967 hCoV-19/England/CAMC-B34299/2020\|EPI_ISL_659541\|20 | ···················· | ···················· | ···················· | ···················· | ···················· | ···················· |
| S | 3968 hCoV-19/England/CAMC-B3467F/2020\|EPI_ISL_659543\|20 | ···················· | ···················· | ···················· | ···················· | ···················· | ···················· |
| S | 3969 hCoV-19/England/CAMC-B32A0C/2020\|EPI_ISL_659544\|20 | ···················· | ···················· | ···················· | ···················· | ···················· | ···················· |
| S | 3970 hCoV-19/England/CAMC-B3431E/2020\|EPI_ISL_659545\|20 | ···················· | ···················· | ···················· | ···················· | ···················· | ···················· |
| S | 3971 hCoV-19/England/CAMC-B357A8/2020\|EPI_ISL_659546\|20 | ···················· | ···················· | ···················· | ···················· | ···················· | ···················· |
| S | 3972 hCoV-19/England/CAMC-B3491C/2020\|EPI_ISL_659547\|20 | ···················· | ···················· | ···················· | ···················· | ···················· | ···················· |
| S | 3973 hCoV-19/England/CAMC-B32AB1/2020\|EPI_ISL_659548\|20 | ···················· | ···················· | ···················· | ···················· | ···················· | ···················· |
| S | 3974 hCoV-19/England/CAMC-B3577B/2020\|EPI_ISL_659550\|20 | ···················· | ···················· | ···················· | ···················· | ···················· | ···················· |
| S | 3975 hCoV-19/England/CAMC-B355EA/2020\|EPI_ISL_659552\|20 | ···················· | ···················· | ···················· | ···················· | ···················· | ···················· |
| S | 3976 hCoV-19/England/CAMC-B34590/2020\|EPI_ISL_659553\|20 | ···················· | ···················· | ···················· | ···················· | ···················· | ···················· |
| S | 3977 hCoV-19/England/CAMC-B32BBE/2020\|EPI_ISL_659554\|20 | ···················· | ···················· | ···················· | ···················· | ···················· | ···················· |
| S | 3978 hCoV-19/England/CAMC-B34563/2020\|EPI_ISL_659555\|20 | ···················· | ···················· | ···················· | ···················· | ······T············· | ···················· |
| S | 3979 hCoV-19/England/CAMC-B356E7/2020\|EPI_ISL_659557\|20 | ···················· | ···················· | ···················· | ···················· | ···················· | ···················· |
| S | 3980 hCoV-19/England/CAMC-B349A3/2020\|EPI_ISL_659560\|20 | ···················· | ···················· | ···················· | ···················· | ···················· | ···················· |
| S | 3981 hCoV-19/England/CAMC-B329C3/2020\|EPI_ISL_659561\|20 | ···················· | ···················· | ···················· | ···················· | ···················· | ···················· |
| S | 3982 hCoV-19/England/CAMC-B34651/2020\|EPI_ISL_659562\|20 | ···················· | ···················· | ···················· | ···················· | ···················· | ···················· |
| S | 3983 hCoV-19/England/CAMC-B34378/2020\|EPI_ISL_659564\|20 | ···················· | ···················· | ···················· | ···················· | ···················· | ···················· |
| S | 3984 hCoV-19/England/CAMC-B34606/2020\|EPI_ISL_659565\|20 | ···················· | ···················· | ···················· | ···················· | ···················· | ···················· |
| S | 3985 hCoV-19/England/CAMC-B327AB/2020\|EPI_ISL_659567\|20 | ···················· | ···················· | ···················· | ···················· | ···················· | ···················· |
| S | 3986 hCoV-19/England/CAMC-B34545/2020\|EPI_ISL_659568\|20 | ···················· | ···················· | ···················· | ···················· | ···················· | ···················· |
| S | 3987 hCoV-19/England/CAMC-B346E8/2020\|EPI_ISL_659570\|20 | ···················· | ···················· | ···················· | ···················· | ···················· | ···················· |
| S | 3988 hCoV-19/England/CAMC-B35395/2020\|EPI_ISL_659572\|20 | ···················· | ···················· | ···················· | ···················· | ···················· | ···················· |
| S | 3989 hCoV-19/England/CAMC-B32ADF/2020\|EPI_ISL_659574\|20 | ···················· | ···················· | ···················· | ···················· | ···················· | ···················· |
| S | 3990 hCoV-19/England/CAMC-B35377/2020\|EPI_ISL_659575\|20 | ···················· | ·A·················· | ···················· | ···················· | ···················· | ··A················· |
| S | 3991 hCoV-19/England/CAMC-B3541A/2020\|EPI_ISL_659578\|20 | ···················· | ···················· | ···················· | ···················· | ···················· | ···················· |
| S | 3992 hCoV-19/England/CAMC-B342B7/2020\|EPI_ISL_659579\|20 | ···················· | ···················· | ···················· | ···················· | ···················· | ···················· |
| S | 3993 hCoV-19/England/CAMC-B32CBB/2020\|EPI_ISL_659580\|20 | ···················· | ·········C·········· | ···················· | ···················· | ···················· | ···················· |
| S | 3994 hCoV-19/England/CAMC-B34B9E/2020\|EPI_ISL_659581\|20 | ···················· | ···················· | ···················· | ···················· | ···················· | ···················· |
| S | 3995 hCoV-19/England/CAMC-B3569C/2020\|EPI_ISL_659582\|20 | ···················· | ···················· | ···················· | ···················· | ···················· | ···················· |
| S | 3996 hCoV-19/England/CAMC-B346CA/2020\|EPI_ISL_659583\|20 | ···················· | ···················· | ···················· | ···················· | ······T············· | ···················· |
| S | 3997 hCoV-19/England/CAMC-B340AE/2020\|EPI_ISL_659584\|20 | ···················· | ···················· | ···················· | ···················· | ···················· | ···················· |
| S | 3998 hCoV-19/England/CAMC-B340CC/2020\|EPI_ISL_659586\|20 | ···················· | ···················· | ···················· | ···················· | ···················· | ···················· |
| S | 3999 hCoV-19/England/CAMC-B32B63/2020\|EPI_ISL_659589\|20 | ···················· | ···················· | ···················· | ···················· | ···················· | ···················· |
| S | 4000 hCoV-19/England/CAMC-B345AF/2020\|EPI_ISL_659591\|20 | ···················· | ···················· | ···················· | ···················· | ···················· | ···················· |
| S | 4001 hCoV-19/England/CAMC-B354A1/2020\|EPI_ISL_659593\|20 | ···················· | ···················· | ···················· | ···················· | ···················· | ···················· |
| S | 4002 hCoV-19/England/CAMC-B3276F/2020\|EPI_ISL_659594\|20 | ···················· | ···················· | ···················· | ···················· | ···················· | ···················· |
| S | 4003 hCoV-19/England/CAMC-B3433C/2020\|EPI_ISL_659596\|20 | ···················· | ···················· | ···················· | ···················· | ···················· | ···················· |
| S | 4004 hCoV-19/England/CAMC-B32D12/2020\|EPI_ISL_659598\|20 | ···················· | ···················· | ···················· | ···················· | ···················· | ···················· |
| S | 4005 hCoV-19/England/CAMC-B3286C/2020\|EPI_ISL_659600\|20 | ···················· | ···················· | ···················· | ···················· | ···················· | ···················· |
| S | 4006 hCoV-19/England/CAMC-B34387/2020\|EPI_ISL_659601\|20 | ···················· | ···················· | ···················· | ···················· | ···················· | ···················· |
| S | 4007 hCoV-19/England/CAMC-B3568D/2020\|EPI_ISL_659602\|20 | ···················· | ···················· | ···················· | ···················· | ···················· | ···················· |
| S | 4008 hCoV-19/England/CAMC-B343F0/2020\|EPI_ISL_659604\|20 | ···················· | ···················· | ···················· | ···················· | ···················· | ···················· |
| S | 4009 hCoV-19/England/CAMC-B35368/2020\|EPI_ISL_659605\|20 | ···················· | ···················· | ···················· | ···················· | ···················· | ···················· |
| S | 4010 hCoV-19/England/CAMC-B34B34/2020\|EPI_ISL_659608\|20 | ···················· | ···················· | ···················· | ···················· | ···················· | ···················· |
| S | 4011 hCoV-19/England/CAMC-B328A8/2020\|EPI_ISL_659610\|20 | ···················· | ···················· | ···················· | ···················· | ···················· | ···················· |
| S | 4012 hCoV-19/England/CAMC-B328B7/2020\|EPI_ISL_659611\|20 | ···················· | ···················· | ···················· | ···················· | ···················· | ···················· |
| S | 4013 hCoV-19/England/CAMC-B347C7/2020\|EPI_ISL_659612\|20 | ···················· | ···················· | ···················· | ···················· | ······T············· | ···················· |
| S | 4014 hCoV-19/England/CAMC-B34615/2020\|EPI_ISL_659613\|20 | ···················· | ···················· | ···················· | ···················· | ···················· | ···················· |

| | | | | | | | |
|---|---|---|---|---|---|---|---|
| S | 4015 hCoV-19/England/CAMC-B349EF/2020\|EPI_ISL_659614\|20 | ···················· | ···················· | ···················· | ···················· | ···················· | ···················· |
| S | 4016 hCoV-19/England/CAMC-B32DD6/2020\|EPI_ISL_659617\|20 | ···················· | ···················· | ···················· | ···················· | ···················· | ···················· |
| S | 4017 hCoV-19/England/CAMC-B328C6/2020\|EPI_ISL_659618\|20 | ···················· | ···················· | ···················· | ···················· | ···················· | ···················· |
| S | 4018 hCoV-19/England/CAMC-B34105/2020\|EPI_ISL_659620\|20 | ···················· | ···················· | ···················· | ···················· | ···················· | ···················· |
| S | 4019 hCoV-19/England/CAMC-B342A8/2020\|EPI_ISL_659622\|20 | ···················· | ···················· | ···················· | ···················· | ···················· | ···················· |
| S | 4020 hCoV-19/England/CAMC-B34A73/2020\|EPI_ISL_659623\|20 | ···················· | ···················· | ···················· | ···················· | ···················· | ···················· |
| S | 4021 hCoV-19/England/CAMC-B34008/2020\|EPI_ISL_659624\|20 | ···················· | ···················· | ···················· | ···················· | ···················· | ···················· |
| S | 4022 hCoV-19/England/CAMC-B34879/2020\|EPI_ISL_659625\|20 | ···················· | ···················· | ···················· | ···················· | ······T············· | ···················· |
| S | 4023 hCoV-19/England/CAMC-B343E1/2020\|EPI_ISL_659626\|20 | ···················· | ···················· | ···················· | ···················· | ···················· | ···················· |
| S | 4024 hCoV-19/England/CAMC-B355DB/2020\|EPI_ISL_659639\|20 | ···················· | ···················· | ···················· | ···················· | ······T············· | ···················· |
| S | 4025 hCoV-19/Ecuador/USFQ-228/2020\|EPI_ISL_660529\|2020- | ···················· | ···················· | ···················· | ···················· | ···················· | ···················· |
| S | 4026 hCoV-19/Ecuador/USFQ-230/2020\|EPI_ISL_660530\|2020- | ···················· | ···················· | ···················· | ···················· | ···················· | ···················· |
| S | 4027 hCoV-19/Ecuador/USFQ-231/2020\|EPI_ISL_660531\|2020- | ···················· | ···················· | ···················· | ···················· | ···················· | ···················· |
| S | 4028 hCoV-19/Ecuador/USFQ-402/2020\|EPI_ISL_660532\|2020- | ···················· | ···················· | ···················· | ···················· | ···················· | ···················· |
| S | 4029 hCoV-19/Ecuador/USFQ-485/2020\|EPI_ISL_660533\|2020- | ···················· | ···················· | ···················· | ···················· | ···················· | ···················· |
| S | 4030 hCoV-19/Ecuador/USFQ-487/2020\|EPI_ISL_660534\|2020- | ···················· | ···················· | ···················· | ···················· | ···················· | ···················· |
| S | 4031 hCoV-19/Ecuador/USFQ-506/2020\|EPI_ISL_660535\|2020- | ···················· | ···················· | ···················· | ···················· | ···················· | ···················· |
| S | 4032 hCoV-19/Ecuador/USFQ-509/2020\|EPI_ISL_660536\|2020- | ···················· | ···················· | ···················· | ···················· | ···················· | ···················· |
| S | 4033 hCoV-19/Ecuador/USFQ-513/2020\|EPI_ISL_660537\|2020- | ···················· | ···················· | ···················· | ···················· | ···················· | ···················· |
| S | 4034 hCoV-19/Ecuador/USFQ-515/2020\|EPI_ISL_660538\|2020- | ···················· | ···················· | ···················· | ···················· | ···················· | ···················· |
| S | 4035 hCoV-19/Ecuador/USFQ-520/2020\|EPI_ISL_660539\|2020- | ···················· | ···················· | ···················· | ···················· | ···················· | ···················· |
| S | 4036 hCoV-19/Vietnam/38238/2020\|EPI_ISL_660541\|2020-01- | ···················· | ···················· | ···················· | ···················· | ···················· | ···················· |
| S | 4037 hCoV-19/Vietnam/39730/2020\|EPI_ISL_660542\|2020-03- | ···················· | ···················· | ···················· | ···················· | ···················· | ···················· |
| S | 4038 hCoV-19/Taiwan/CGMH-CGU-31/2020\|EPI_ISL_660543\|202 | ···················· | ···················· | ···················· | ···················· | ···················· | ···················· |
| S | 4039 hCoV-19/Taiwan/CGMH-CGU-32/2020\|EPI_ISL_660544\|202 | ···················· | ···················· | ···················· | ···················· | ······T············· | ···················· |
| S | 4040 hCoV-19/Taiwan/CGMH-CGU-33/2020\|EPI_ISL_660545\|202 | ···················· | ···················· | ···················· | ···················· | ···················· | ···················· |
| S | 4041 hCoV-19/England/204380390/2020\|EPI_ISL_660547\|2020 | ···················· | ···················· | ···················· | ···················· | ···················· | ···················· |
| S | 4042 hCoV-19/England/204380637/2020\|EPI_ISL_660548\|2020 | ···················· | ···················· | ···················· | ···················· | ···················· | ···················· |
| S | 4043 hCoV-19/England/204390705/2020\|EPI_ISL_660549\|2020 | ···················· | ···················· | ···················· | ···················· | ···················· | ···················· |
| S | 4044 hCoV-19/Czech Republic/NRL-10233/2020\|EPI_ISL_6605 | ···················· | ···················· | ···················· | ···················· | ···················· | ···················· |
| S | 4045 hCoV-19/Czech Republic/NRL-9962/2020\|EPI_ISL_66055 | ···················· | ···················· | ···················· | ···················· | ···················· | ···················· |
| S | 4046 hCoV-19/Czech Republic/NRL-10281/2020\|EPI_ISL_6605 | ···················· | ······C············· | ···················· | ···················· | ···················· | ···················· |
| S | 4047 hCoV-19/Czech Republic/NRL-10242/2020\|EPI_ISL_6605 | ···················· | ···················· | ···················· | ···················· | ···················· | ···················· |
| S | 4048 hCoV-19/Czech Republic/NRL-10288/2020\|EPI_ISL_6605 | ···················· | ···················· | ···················· | ···················· | ···················· | ···················· |
| S | 4049 hCoV-19/Czech Republic/NRL-9965/2020\|EPI_ISL_66055 | ···················· | ···················· | ···················· | ···················· | ···················· | ···················· |
| S | 4050 hCoV-19/Czech Republic/NRL-9969/2020\|EPI_ISL_66055 | ···················· | ···················· | ···················· | ···················· | ···················· | ···················· |
| S | 4051 hCoV-19/Czech Republic/NRL-9990/2020\|EPI_ISL_66056 | ···················· | ···················· | ···················· | ···················· | ···················· | ···················· |
| S | 4052 hCoV-19/Czech Republic/NRL-10015/2020\|EPI_ISL_6605 | ···················· | ···················· | ···················· | ···················· | ···················· | ···················· |
| S | 4053 hCoV-19/Czech Republic/NRL-10020/2020\|EPI_ISL_6605 | ···················· | ···················· | ···················· | ···················· | ···················· | ···················· |
| S | 4054 hCoV-19/Czech Republic/NRL-10317/2020\|EPI_ISL_6605 | ···················· | ···················· | ···················· | ···················· | ···················· | ···················· |
| S | 4055 hCoV-19/Czech Republic/NRL-9904/2020\|EPI_ISL_66056 | ···················· | ···················· | ···················· | ···················· | ···················· | ···················· |
| S | 4056 hCoV-19/Czech Republic/NRL-10295/2020\|EPI_ISL_6605 | ···················· | ···················· | ···················· | ···················· | ···················· | ···················· |
| S | 4057 hCoV-19/Czech Republic/NRL-10176/2020\|EPI_ISL_6605 | ···················· | ···················· | ···················· | ···················· | ···················· | ···················· |
| S | 4058 hCoV-19/Czech Republic/NRL-10114/2020\|EPI_ISL_6605 | ···················· | ···················· | ···················· | ···················· | ···················· | ···················· |
| S | 4059 hCoV-19/Czech Republic/NRL-10294/2020\|EPI_ISL_6605 | ···················· | ···················· | ···················· | ···················· | ···················· | ···················· |
| S | 4060 hCoV-19/Czech Republic/NRL-10126/2020\|EPI_ISL_6605 | ···················· | ···················· | ···················· | ···················· | ···················· | ···················· |
| S | 4061 hCoV-19/Czech Republic/NRL-10092/2020\|EPI_ISL_6605 | ···················· | ···················· | ···················· | ···················· | ···················· | ···················· |
| S | 4062 hCoV-19/Czech Republic/NRL-10145/2020\|EPI_ISL_6605 | ···················· | ···················· | ···················· | ···················· | ···················· | ···················· |
| S | 4063 hCoV-19/Czech Republic/NRL-10197/2020\|EPI_ISL_6605 | ···················· | ···················· | ···················· | ···················· | ···················· | ···················· |
| S | 4064 hCoV-19/Czech Republic/NRL-10207/2020\|EPI_ISL_6605 | ···················· | ···················· | ···················· | ···················· | ···················· | ···················· |
| S | 4065 hCoV-19/Czech Republic/NRL-10330/2020\|EPI_ISL_6605 | ···················· | ···················· | ···················· | ···················· | ···················· | ···················· |
| S | 4066 hCoV-19/Czech Republic/NRL-10143/2020\|EPI_ISL_6605 | ···················· | ···················· | ···················· | ···················· | ···················· | ···················· |
| S | 4067 hCoV-19/Czech Republic/NRL-10269/2020\|EPI_ISL_6605 | ···················· | ···················· | ···················· | ···················· | ···················· | ···················· |
| S | 4068 hCoV-19/Czech Republic/NRL-10056/2020\|EPI_ISL_6605 | ···················· | ···················· | ···················· | ···················· | ···················· | ···················· |
| S | 4069 hCoV-19/Czech Republic/NRL-10227/2020\|EPI_ISL_6605 | ···················· | ···················· | ···················· | ···················· | ···················· | ···················· |
| S | 4070 hCoV-19/Czech Republic/NRL-10088/2020\|EPI_ISL_6605 | ···················· | ···················· | ···················· | ···················· | ···················· | ···················· |
| S | 4071 hCoV-19/Czech Republic/NRL-10208/2020\|EPI_ISL_6605 | ···················· | ···················· | ···················· | ···················· | ···················· | ···················· |
| S | 4072 hCoV-19/Czech Republic/NRL-10121/2020\|EPI_ISL_6605 | ···················· | ···················· | ···················· | ···················· | ···················· | ···················· |
| S | 4073 hCoV-19/Czech Republic/NRL-10151/2020\|EPI_ISL_6605 | ···················· | ···················· | ···················· | ···················· | ···················· | ···················· |
| S | 4074 hCoV-19/Czech Republic/NRL-10213/2020\|EPI_ISL_6605 | ···················· | ···················· | ···················· | ···················· | ···················· | ···················· |
| S | 4075 hCoV-19/Czech Republic/NRL-10218/2020\|EPI_ISL_6605 | ···················· | ···················· | ···················· | ···················· | ···················· | ···················· |
| S | 4076 hCoV-19/Czech Republic/NRL-10225/2020\|EPI_ISL_6605 | ···················· | ···················· | ···················· | ···················· | ···················· | ···················· |
| S | 4077 hCoV-19/Czech Republic/NRL-9908/2020\|EPI_ISL_66058 | ···················· | ···················· | ···················· | ···················· | ···················· | ···················· |
| S | 4078 hCoV-19/Czech Republic/NRL-9939/2020\|EPI_ISL_66059 | ···················· | ···················· | ···················· | ···················· | ···················· | ···················· |
| S | 4079 hCoV-19/Czech Republic/NRL-10244/2020\|EPI_ISL_6605 | ···················· | ···················· | ···················· | ···················· | ···················· | ···················· |
| S | 4080 hCoV-19/Czech Republic/NRL-10324/2020\|EPI_ISL_6605 | ···················· | ···················· | ···················· | ···················· | ···················· | ···················· |
| S | 4081 hCoV-19/Czech Republic/NRL-10326/2020\|EPI_ISL_6605 | ···················· | ···················· | ···················· | ···················· | ···················· | ···················· |
| S | 4082 hCoV-19/Czech Republic/NRL-10273/2020\|EPI_ISL_6605 | ···················· | ···················· | ···················· | ···················· | ···················· | ···················· |
| S | 4083 hCoV-19/England/204394007/2020\|EPI_ISL_660601\|2020 | ···················· | ···················· | ···················· | ···················· | ···················· | ···················· |
| S | 4084 hCoV-19/England/204441634/2020\|EPI_ISL_660604\|2020 | ···················· | ···················· | ···················· | ···················· | ···················· | ···················· |
| S | 4088 hCoV-19/South Africa/KRISP-K004590/2020\|EPI_ISL_66 | ···················· | ···················· | ···T················ | ···················· | ···················· | ···················· |
| S | 4089 hCoV-19/South Africa/KRISP-K004591/2020\|EPI_ISL_66 | ···················· | ···················· | ···················· | ···················· | ···················· | ···················· |
| S | 4090 hCoV-19/South Africa/KRISP-K004592/2020\|EPI_ISL_66 | ···················· | ···················· | ···················· | ···················· | ···················· | ···················· |
| S | 4091 hCoV-19/South Africa/KRISP-K004593/2020\|EPI_ISL_66 | ···················· | ···················· | ···················· | ···················· | ···················· | ···················· |
| S | 4092 hCoV-19/South Africa/KRISP-K004594/2020\|EPI_ISL_66 | ···················· | ···················· | ···················· | ···················· | ···················· | ···················· |
| S | 4093 hCoV-19/South Africa/KRISP-K004595/2020\|EPI_ISL_66 | ···················· | ·A·················· | ···················· | ···················· | ···················· | ···················· |
| S | 4094 hCoV-19/South Africa/KRISP-K004596/2020\|EPI_ISL_66 | ···················· | ·A·················· | ···················· | ···················· | ···················· | ···················· |
| S | 4095 hCoV-19/South Africa/KRISP-K004597/2020\|EPI_ISL_66 | ···················· | ···················· | ···················· | ···················· | ···················· | ···················· |
| S | 4096 hCoV-19/South Africa/KRISP-K004598/2020\|EPI_ISL_66 | ···················· | ···················· | ···················· | ···················· | ···················· | ···················· |
| S | 4097 hCoV-19/Scotland/EDB8804/2020\|EPI_ISL_652000\|2020- | ···················· | ···················· | ···················· | ···················· | ···················· | ···················· |

| | | | | | | | |
|---|---|---|---|---|---|---|---|
| S | 4098 hCoV-19/Scotland/EDB8813/2020\|EPI_ISL_652001\|2020- | ···················· | ···················· | ····················· | ···················· | ···················· | ···················· |
| S | 4099 hCoV-19/Scotland/EDB8814/2020\|EPI_ISL_652002\|2020- | ···················· | ··········A········· | ····················· | ···················· | ···················· | ···················· |
| S | 4100 hCoV-19/Scotland/EDB8818/2020\|EPI_ISL_652003\|2020- | ···················· | ···················· | ····················· | ···················· | ···················· | ···················· |
| S | 4101 hCoV-19/Scotland/EDB8820/2020\|EPI_ISL_652004\|2020- | ···················· | ···················· | ····················· | ···················· | ···················· | ···················· |
| S | 4102 hCoV-19/Scotland/EDB8825/2020\|EPI_ISL_652005\|2020- | ···················· | ···················· | ····················· | ···················· | ···················· | ···················· |
| S | 4103 hCoV-19/Scotland/EDB8826/2020\|EPI_ISL_652006\|2020- | ···················· | ···················· | ····················· | ···················· | ···················· | ···················· |
| S | 4104 hCoV-19/Scotland/EDB8831/2020\|EPI_ISL_652007\|2020- | ···················· | ···················· | ····················· | ···················· | ···················· | ···················· |
| S | 4105 hCoV-19/Scotland/EDB8836/2020\|EPI_ISL_652008\|2020- | ···················· | ···················· | ····················· | ···················· | ···················· | ···················· |
| S | 4106 hCoV-19/Scotland/EDB8838/2020\|EPI_ISL_652009\|2020- | ···················· | ···················· | ····················· | ···················· | ···················· | ···················· |
| S | 4107 hCoV-19/Scotland/EDB8866/2020\|EPI_ISL_652010\|2020- | ···················· | ···················· | ····················· | ···················· | ···················· | ···················· |
| S | 4108 hCoV-19/Scotland/EDB8868/2020\|EPI_ISL_652011\|2020- | ···················· | ···················· | ····················· | ···················· | ···················· | ···················· |
| S | 4110 hCoV-19/Scotland/EDB8872/2020\|EPI_ISL_652013\|2020- | ···················· | ···················· | ····················· | ···················· | ···················· | ···················· |
| S | 4111 hCoV-19/Scotland/EDB8877/2020\|EPI_ISL_652014\|2020- | ···················· | ···················· | ····················· | ···················· | ···················· | ···················· |
| S | 4112 hCoV-19/Scotland/EDB8881/2020\|EPI_ISL_652015\|2020- | ···················· | ···················· | ····················· | ···················· | ···················· | ···················· |
| S | 4113 hCoV-19/Scotland/EDB8884/2020\|EPI_ISL_652016\|2020- | ···················· | ···················· | ····················· | ···················· | ···················· | ···················· |
| S | 4114 hCoV-19/Scotland/EDB8886/2020\|EPI_ISL_652017\|2020- | ···················· | ···················· | ····················· | ···················· | ···················· | ···················· |
| S | 4115 hCoV-19/Scotland/EDB8897/2020\|EPI_ISL_652018\|2020- | ···················· | ···················· | ····················· | ···················· | ···················· | ···················· |
| S | 4116 hCoV-19/Scotland/EDB8898/2020\|EPI_ISL_652019\|2020- | ···················· | ···················· | ····················· | ···················· | ···················· | ···················· |
| S | 4119 hCoV-19/Scotland/EDB8933/2020\|EPI_ISL_652022\|2020- | ···················· | ···················· | ····················· | ···················· | ···················· | ···················· |
| S | 4120 hCoV-19/Scotland/EDB8957/2020\|EPI_ISL_652023\|2020- | ···················· | ···················· | ····················· | ···················· | ···················· | ···················· |
| S | 4121 hCoV-19/Scotland/EDB8967/2020\|EPI_ISL_652024\|2020- | ···················· | ···················· | ····················· | ···················· | ···················· | ···················· |
| S | 4127 hCoV-19/Scotland/EDB8993/2020\|EPI_ISL_652030\|2020- | ···················· | ···················· | ····················· | ···················· | ···················· | ···················· |
| S | 4128 hCoV-19/Scotland/EDB8994/2020\|EPI_ISL_652031\|2020- | ···················· | ···················· | ····················· | ···················· | ···················· | ···················· |
| S | 4129 hCoV-19/Scotland/EDB9006/2020\|EPI_ISL_652032\|2020- | ···················· | ···················· | ····················· | ···················· | ···················· | ···················· |
| S | 4132 hCoV-19/Scotland/EDB9031/2020\|EPI_ISL_652035\|2020- | ···················· | ···················· | ····················· | ···················· | ···················· | ···················· |
| S | 4133 hCoV-19/Scotland/EDB9033/2020\|EPI_ISL_652036\|2020- | ···················· | ···················· | ····················· | ···················· | ···················· | ···················· |
| S | 4134 hCoV-19/Scotland/EDB9040/2020\|EPI_ISL_652037\|2020- | ···················· | ···················· | ····················· | ···················· | ···················· | ···················· |
| S | 4138 hCoV-19/Scotland/EDB9105/2020\|EPI_ISL_652041\|2020- | ···················· | ···················· | ····················· | ···················· | ···················· | ···················· |
| S | 4139 hCoV-19/Scotland/EDB9107/2020\|EPI_ISL_652042\|2020- | ···················· | ···················· | ····················· | ···················· | ···················· | ···················· |
| S | 4140 hCoV-19/Scotland/EDB9108/2020\|EPI_ISL_652043\|2020- | ···················· | ···················· | ····················· | ···················· | ···················· | ···················· |
| S | 4141 hCoV-19/Scotland/EDB9109/2020\|EPI_ISL_652044\|2020- | ···················· | ···················· | ····················· | ···················· | ···················· | ···················· |
| S | 4142 hCoV-19/Scotland/EDB9110/2020\|EPI_ISL_652045\|2020- | ···················· | ···················· | ····················· | ···················· | ···················· | ···················· |
| S | 4143 hCoV-19/Scotland/EDB9111/2020\|EPI_ISL_652046\|2020- | ···················· | ···················· | ····················· | ···················· | ···················· | ···················· |
| S | 4144 hCoV-19/Scotland/EDB9112/2020\|EPI_ISL_652047\|2020- | ···················· | ···················· | ····················· | ···················· | ···················· | ···················· |
| S | 4145 hCoV-19/Scotland/EDB9115/2020\|EPI_ISL_652048\|2020- | ···················· | ···················· | ····················· | ···················· | ···················· | ···················· |
| S | 4146 hCoV-19/Scotland/EDB9206/2020\|EPI_ISL_652049\|2020- | ···················· | ···················· | ····················· | ···················· | ···················· | ···················· |
| S | 4147 hCoV-19/Scotland/EDB9221/2020\|EPI_ISL_652050\|2020- | ···················· | ···················· | ····················· | ···················· | ···················· | ···················· |
| S | 4149 hCoV-19/Scotland/EDB9243/2020\|EPI_ISL_652052\|2020- | ···················· | ···················· | ····················· | ···················· | ···················· | ···················· |
| S | 4150 hCoV-19/Scotland/EDB9253/2020\|EPI_ISL_652053\|2020- | ···················· | ···················· | ····················· | ···················· | ···················· | ···················· |
| S | 4151 hCoV-19/Scotland/EDB9256/2020\|EPI_ISL_652054\|2020- | ···················· | ···················· | ····················· | ···················· | ···················· | ···················· |
| S | 4152 hCoV-19/Scotland/EDB9257/2020\|EPI_ISL_652055\|2020- | ···················· | ···················· | ····················· | ···················· | ···················· | ···················· |
| S | 4153 hCoV-19/Scotland/EDB9322/2020\|EPI_ISL_652056\|2020- | ···················· | ···················· | ····················· | ···················· | ···················· | ···················· |
| S | 4154 hCoV-19/Scotland/EDB9453/2020\|EPI_ISL_652057\|2020- | ···················· | ···················· | ····················· | ···················· | ···················· | ···················· |
| S | 4156 hCoV-19/Scotland/EDB9458/2020\|EPI_ISL_652059\|2020- | ···················· | ···················· | ····················· | ···················· | ···················· | ···················· |
| S | 4157 hCoV-19/USA/FL-BPHL-2150/2020\|EPI_ISL_653303\|2020- | ···················· | ···················· | ····················· | ···················· | ···················· | ···················· |
| S | 4158 hCoV-19/USA/FL-BPHL-2151/2020\|EPI_ISL_653304\|2020- | ···················· | ···················· | ····················· | ···················· | ···················· | ···················· |
| S | 4159 hCoV-19/USA/FL-BPHL-2153/2020\|EPI_ISL_653306\|2020- | ···················· | ···················· | ····················· | ···················· | ···················· | ···················· |
| S | 4160 hCoV-19/USA/FL-BPHL-2164/2020\|EPI_ISL_653317\|2020- | ···················· | ···················· | ····················· | ···················· | ···················· | ···················· |
| S | 4161 hCoV-19/USA/FL-BPHL-2167/2020\|EPI_ISL_653320\|2020- | ···················· | ···················· | ····················· | ···················· | ···················· | ···················· |
| S | 4162 hCoV-19/USA/FL-BPHL-2168/2020\|EPI_ISL_653321\|2020- | ···················· | ···················· | ····················· | ···················· | ···················· | ···················· |
| S | 4163 hCoV-19/USA/FL-BPHL-2170/2020\|EPI_ISL_653323\|2020- | ···················· | ···················· | ····················· | ···················· | ···················· | ···················· |
| S | 4164 hCoV-19/USA/FL-BPHL-2171/2020\|EPI_ISL_653324\|2020- | ···················· | ···················· | ····················· | ···················· | ···················· | ···················· |
| S | 4165 hCoV-19/USA/FL-BPHL-2173/2020\|EPI_ISL_653326\|2020- | ···················· | ···················· | ····················· | ···················· | ···················· | ···················· |
| S | 4166 hCoV-19/USA/FL-BPHL-2176/2020\|EPI_ISL_653329\|2020- | ···················· | ···················· | ····················· | ···················· | ···················· | ···················· |
| S | 4167 hCoV-19/USA/FL-BPHL-2178/2020\|EPI_ISL_653331\|2020- | ···················· | ···················· | ····················· | ···················· | ···················· | ···················· |
| S | 4169 hCoV-19/USA/FL-BPHL-2186/2020\|EPI_ISL_653339\|2020- | ···················· | ···················· | ····················· | ···················· | ···················· | ···················· |
| S | 4170 hCoV-19/USA/FL-BPHL-2188/2020\|EPI_ISL_653341\|2020- | ···················· | ···················· | ····················· | ···················· | ···················· | ···················· |
| S | 4171 hCoV-19/USA/LA-EVTL654/2020\|EPI_ISL_653346\|2020-07 | ···················· | ···················· | ····················· | ···················· | ···················· | ···················· |
| S | 4172 hCoV-19/USA/LA-EVTL655/2020\|EPI_ISL_653347\|2020-07 | ···················· | ···················· | ····················· | ···················· | ···················· | ···················· |
| S | 4173 hCoV-19/USA/LA-EVTL656/2020\|EPI_ISL_653348\|2020-07 | ···················· | ···················· | ····················· | ···················· | ···················· | ···················· |
| S | 4174 hCoV-19/USA/LA-EVTL657/2020\|EPI_ISL_653349\|2020-07 | ···················· | ···················· | ····················· | ···················· | ···················· | ···················· |
| S | 4175 hCoV-19/USA/LA-EVTL658/2020\|EPI_ISL_653350\|2020-07 | ···················· | ···················· | ····················· | ···················· | ···················· | ···················· |
| S | 4176 hCoV-19/USA/LA-EVTL659/2020\|EPI_ISL_653351\|2020-07 | ···················· | ···················· | ····················· | ···················· | ···················· | ···················· |
| S | 4177 hCoV-19/USA/LA-EVTL660/2020\|EPI_ISL_653352\|2020-07 | ···················· | ···················· | ····················· | ···················· | ···················· | ···················· |
| S | 4178 hCoV-19/USA/LA-EVTL661/2020\|EPI_ISL_653353\|2020-07 | ···················· | ···················· | ····················· | ···················· | ···················· | ···················· |
| S | 4179 hCoV-19/USA/LA-EVTL662/2020\|EPI_ISL_653354\|2020-07 | ···················· | ···················· | ····················· | ···················· | ···················· | ···················· |
| S | 4180 hCoV-19/USA/LA-EVTL663/2020\|EPI_ISL_653355\|2020-07 | ···················· | ···················· | ····················· | ···················· | ···················· | ···················· |
| S | 4181 hCoV-19/USA/LA-EVTL664/2020\|EPI_ISL_653356\|2020-07 | ···················· | ···················· | ····················· | ···················· | ···················· | ···················· |
| S | 4182 hCoV-19/USA/LA-EVTL665/2020\|EPI_ISL_653357\|2020-07 | ···················· | ···················· | ····················· | ···················· | ···················· | ···················· |
| S | 4183 hCoV-19/USA/LA-EVTL666/2020\|EPI_ISL_653358\|2020-07 | ···················· | ···················· | ····················· | ···················· | ···················· | ···················· |
| S | 4184 hCoV-19/USA/LA-EVTL667/2020\|EPI_ISL_653359\|2020-07 | ···················· | ···················· | ····················· | ···················· | ···················· | ···················· |
| S | 4185 hCoV-19/USA/LA-EVTL668/2020\|EPI_ISL_653360\|2020-07 | ···················· | ···················· | ····················· | ···················· | ···················· | ···················· |
| S | 4186 hCoV-19/USA/LA-EVTL669/2020\|EPI_ISL_653361\|2020-07 | ···················· | ···················· | ····················· | ···················· | ···················· | ···················· |
| S | 4187 hCoV-19/USA/LA-EVTL670/2020\|EPI_ISL_653362\|2020-07 | ···················· | ···················· | ····················· | ···················· | ···················· | ···················· |
| S | 4188 hCoV-19/USA/LA-EVTL671/2020\|EPI_ISL_653363\|2020-07 | ···················· | ···················· | ····················· | ···················· | ···················· | ···················· |
| S | 4189 hCoV-19/USA/LA-EVTL672/2020\|EPI_ISL_653364\|2020-07 | ···················· | ···················· | ····················· | ···················· | ···················· | ···················· |
| S | 4190 hCoV-19/USA/LA-EVTL673/2020\|EPI_ISL_653365\|2020-07 | ···················· | ·A·················· | ····················· | ···················· | ···················· | ···················· |
| S | 4191 hCoV-19/USA/LA-EVTL674/2020\|EPI_ISL_653366\|2020-07 | ···················· | ···················· | ····················· | ···················· | ···················· | ···················· |
| S | 4192 hCoV-19/USA/LA-EVTL676/2020\|EPI_ISL_653368\|2020-07 | ···················· | ···················· | ····················· | ···················· | ···················· | ···················· |
| S | 4193 hCoV-19/USA/LA-EVTL677/2020\|EPI_ISL_653369\|2020-07 | ···················· | ···················· | ····················· | ···················· | ···················· | ···················· |

| | | | | | | | |
|---|---|---|---|---|---|---|---|
| S | 4194 hCoV-19/USA/LA-EVTL678/2020\|EPI_ISL_653370\|2020-07 | ···················· | ······A············· | ···················· | ···················· | ···················· | ···················· |
| S | 4195 hCoV-19/USA/LA-EVTL679/2020\|EPI_ISL_653371\|2020-07 | ···················· | ······A············· | ···················· | ···················· | ···················· | ···················· |
| S | 4196 hCoV-19/USA/LA-EVTL680/2020\|EPI_ISL_653372\|2020-07 | ···················· | ······A············· | ···················· | ···················· | ···················· | ···················· |
| S | 4197 hCoV-19/USA/LA-EVTL681/2020\|EPI_ISL_653373\|2020-07 | ···················· | ······A············· | ···················· | ···················· | ···················· | ···················· |
| S | 4198 hCoV-19/USA/LA-EVTL682/2020\|EPI_ISL_653374\|2020-07 | ···················· | ······A············· | ···················· | ···················· | ···················· | ···················· |
| S | 4199 hCoV-19/USA/LA-EVTL683/2020\|EPI_ISL_653375\|2020-07 | ···················· | ·A·················· | ···················· | ···················· | ···················· | ···················· |
| S | 4200 hCoV-19/USA/LA-EVTL684/2020\|EPI_ISL_653376\|2020-07 | ···················· | ···················· | ···················· | ···················· | ···················· | ···················· |
| S | 4201 hCoV-19/USA/LA-EVTL685/2020\|EPI_ISL_653377\|2020-07 | ···················· | ···················· | ···················· | ···················· | ···················· | ···················· |
| S | 4202 hCoV-19/USA/LA-EVTL686/2020\|EPI_ISL_653378\|2020-07 | ···················· | ···················· | ···················· | ···················· | ···················· | ···················· |
| S | 4203 hCoV-19/USA/LA-EVTL687/2020\|EPI_ISL_653379\|2020-07 | ···················· | ·A·················· | ···················· | ···················· | ···················· | ···················· |
| S | 4204 hCoV-19/USA/LA-EVTL688/2020\|EPI_ISL_653380\|2020-07 | ···················· | ···················· | ···················· | ···················· | ···················· | ···················· |
| S | 4205 hCoV-19/USA/LA-EVTL689/2020\|EPI_ISL_653381\|2020-07 | ···················· | ···················· | ···················· | ···················· | ···················· | ···················· |
| S | 4206 hCoV-19/USA/LA-EVTL690/2020\|EPI_ISL_653382\|2020-07 | ···················· | ···················· | ···················· | ···················· | ···················· | ···················· |
| S | 4207 hCoV-19/USA/LA-EVTL691/2020\|EPI_ISL_653383\|2020-07 | ···················· | ···················· | ···················· | ···················· | ···················· | ···················· |
| S | 4208 hCoV-19/USA/LA-EVTL692/2020\|EPI_ISL_653384\|2020-07 | ···················· | ···················· | ···················· | ···················· | ···················· | ···················· |
| S | 4209 hCoV-19/USA/LA-EVTL693/2020\|EPI_ISL_653385\|2020-07 | ···················· | ···················· | ···················· | ···················· | ···················· | ···················· |
| S | 4210 hCoV-19/USA/LA-EVTL694/2020\|EPI_ISL_653386\|2020-07 | ···················· | ···················· | ···················· | ···················· | ···················· | ···················· |
| S | 4211 hCoV-19/USA/LA-EVTL695/2020\|EPI_ISL_653387\|2020-07 | ···················· | ···················· | ···················· | ···················· | ···················· | ···················· |
| S | 4212 hCoV-19/USA/LA-EVTL696/2020\|EPI_ISL_653388\|2020-07 | ···················· | ···················· | ···················· | ···················· | ···················· | ···················· |
| S | 4213 hCoV-19/USA/LA-EVTL697/2020\|EPI_ISL_653389\|2020-07 | ···················· | ···················· | ···················· | ···················· | ···················· | ···················· |
| S | 4214 hCoV-19/USA/LA-EVTL708/2020\|EPI_ISL_653400\|2020-07 | ···················· | ···················· | ···················· | ···················· | ···················· | ···················· |
| S | 4215 hCoV-19/USA/LA-EVTL709/2020\|EPI_ISL_653401\|2020-07 | ···················· | ···················· | ···················· | ···················· | ···················· | ···················· |
| S | 4216 hCoV-19/Spain/AS-IBV-98006648/2020\|EPI_ISL_654634\| | ···················· | ···················· | ···················· | ···················· | ···················· | ···················· |
| S | 4219 hCoV-19/Spain/AS-IBV-98006589/2020\|EPI_ISL_654637\| | ···················· | ···················· | ···················· | ···················· | ···················· | ···················· |
| S | 4220 hCoV-19/Spain/AS-IBV-98006592/2020\|EPI_ISL_654638\| | ···················· | ···················· | ···················· | ···················· | ···················· | ···················· |
| S | 4221 hCoV-19/Spain/AS-IBV-98006594/2020\|EPI_ISL_654639\| | ···················· | ···················· | ···················· | ···················· | ···················· | ···················· |
| S | 4222 hCoV-19/Spain/AS-IBV-98006595/2020\|EPI_ISL_654640\| | ···················· | ···················· | ···················· | ···················· | ···················· | ···················· |
| S | 4223 hCoV-19/Spain/AS-IBV-98006597/2020\|EPI_ISL_654641\| | ···················· | ···················· | ···················· | ···················· | ···················· | ···················· |
| S | 4224 hCoV-19/Spain/AS-IBV-98006598/2020\|EPI_ISL_654642\| | ···················· | ···················· | ···················· | ···················· | ···················· | ···················· |
| S | 4225 hCoV-19/Spain/AS-IBV-98006599/2020\|EPI_ISL_654643\| | ···················· | ···················· | ···················· | ···················· | ···················· | ···················· |
| S | 4226 hCoV-19/Spain/AS-IBV-98006600/2020\|EPI_ISL_654644\| | ···················· | ···················· | ···················· | ···················· | ···················· | ···················· |
| S | 4227 hCoV-19/Spain/AS-IBV-98006601/2020\|EPI_ISL_654645\| | ···················· | ···················· | ···················· | ···················· | ···················· | ···················· |
| S | 4228 hCoV-19/Spain/AS-IBV-98006602/2020\|EPI_ISL_654646\| | ···················· | ···················· | ···················· | ···················· | ···················· | ···················· |
| S | 4229 hCoV-19/Spain/AS-IBV-98006603/2020\|EPI_ISL_654647\| | ···················· | ···················· | ···················· | ···················· | ···················· | ···················· |
| S | 4230 hCoV-19/Spain/AS-IBV-98006604/2020\|EPI_ISL_654648\| | ···················· | ···················· | ···················· | ···················· | ···················· | ···················· |
| S | 4231 hCoV-19/Spain/AS-IBV-98006605/2020\|EPI_ISL_654649\| | ···················· | ···················· | ···················· | ···················· | ···················· | ···················· |
| S | 4232 hCoV-19/Spain/AS-IBV-98006606/2020\|EPI_ISL_654650\| | ···················· | ···················· | ···················· | ···················· | ···················· | ···················· |
| S | 4233 hCoV-19/Spain/AS-IBV-98006607/2020\|EPI_ISL_654651\| | ···················· | ···················· | ···················· | ···················· | ···················· | ···················· |
| S | 4234 hCoV-19/Spain/AS-IBV-98006608/2020\|EPI_ISL_654652\| | ···················· | ···················· | ···················· | ···················· | ···················· | ···················· |
| S | 4235 hCoV-19/Spain/AS-IBV-98006610/2020\|EPI_ISL_654653\| | ···················· | ···················· | ···················· | ···················· | ···················· | ···················· |
| S | 4236 hCoV-19/Spain/AS-IBV-98006611/2020\|EPI_ISL_654654\| | ···················· | ···················· | ···················· | ···················· | ···················· | ···················· |
| S | 4237 hCoV-19/Spain/AS-IBV-98006612/2020\|EPI_ISL_654655\| | ···················· | ···················· | ···················· | ···················· | ···················· | ···················· |
| S | 4238 hCoV-19/Spain/AS-IBV-98006613/2020\|EPI_ISL_654656\| | ···················· | ···················· | ···················· | ···················· | ···················· | ···················· |
| S | 4239 hCoV-19/Spain/AS-IBV-98006616/2020\|EPI_ISL_654657\| | ···················· | ···················· | ···················· | ···················· | ···················· | ···················· |
| S | 4240 hCoV-19/Spain/AS-IBV-98006618/2020\|EPI_ISL_654658\| | ···················· | ···················· | ···················· | ···················· | ···················· | ···················· |
| S | 4241 hCoV-19/Spain/AS-IBV-98006620/2020\|EPI_ISL_654659\| | ···················· | ···················· | ···················· | ···················· | ···················· | ···················· |
| S | 4242 hCoV-19/Spain/AS-IBV-98006623/2020\|EPI_ISL_654660\| | ···················· | ···················· | ···················· | ···················· | ···················· | ···················· |
| S | 4243 hCoV-19/Spain/AS-IBV-98006625/2020\|EPI_ISL_654661\| | ···················· | ···················· | ···················· | ···················· | ···················· | ···················· |
| S | 4244 hCoV-19/Spain/AS-IBV-98006626/2020\|EPI_ISL_654662\| | ···················· | ···················· | ···················· | ···················· | ···················· | ···················· |
| S | 4245 hCoV-19/Spain/AS-IBV-98006629/2020\|EPI_ISL_654663\| | ···················· | ···················· | ···················· | ···················· | ···················· | ···················· |
| S | 4246 hCoV-19/Spain/AS-IBV-98006630/2020\|EPI_ISL_654664\| | ···················· | ···················· | ···················· | ···················· | ···················· | ···················· |
| S | 4247 hCoV-19/Spain/AS-IBV-98006631/2020\|EPI_ISL_654665\| | ···················· | ···················· | ···················· | ···················· | ···················· | ···················· |
| S | 4248 hCoV-19/Spain/AS-IBV-98006632/2020\|EPI_ISL_654666\| | ···················· | ···················· | ···················· | ···················· | ···················· | ···················· |
| S | 4249 hCoV-19/Spain/AS-IBV-98006634/2020\|EPI_ISL_654667\| | ···················· | ···················· | ···················· | ···················· | ···················· | ···················· |
| S | 4250 hCoV-19/Spain/AS-IBV-98006638/2020\|EPI_ISL_654668\| | ···················· | ···················· | ···················· | ···················· | ···················· | ···················· |
| S | 4251 hCoV-19/Spain/AS-IBV-98006643/2020\|EPI_ISL_654669\| | ···················· | ···················· | ···················· | ···················· | ···················· | ···················· |
| S | 4252 hCoV-19/Spain/AS-IBV-98006644/2020\|EPI_ISL_654670\| | ···················· | ···················· | ···················· | ···················· | ···················· | ···················· |
| S | 4253 hCoV-19/Spain/AS-IBV-98006656/2020\|EPI_ISL_654671\| | ···················· | ···················· | ···················· | ···················· | ···················· | ···················· |
| S | 4254 hCoV-19/Spain/AS-IBV-98006615/2020\|EPI_ISL_654672\| | ···················· | ···················· | ···················· | ···················· | ···················· | ···················· |
| S | 4255 hCoV-19/Spain/AS-IBV-98006617/2020\|EPI_ISL_654673\| | ···················· | ···················· | ···················· | ···················· | ···················· | ···················· |
| S | 4256 hCoV-19/Spain/AS-IBV-98006650/2020\|EPI_ISL_654674\| | ···················· | ···················· | ···················· | ···················· | ···················· | ···················· |
| S | 4257 hCoV-19/Spain/AS-IBV-98006621/2020\|EPI_ISL_654675\| | ···················· | ···················· | ···················· | ···················· | ···················· | ···················· |
| S | 4258 hCoV-19/Spain/AS-IBV-98006635/2020\|EPI_ISL_654676\| | ···················· | ···················· | ···················· | ···················· | ···················· | ···················· |
| S | 4259 hCoV-19/Spain/AS-IBV-98006636/2020\|EPI_ISL_654677\| | ···················· | ···················· | ···················· | ···················· | ···················· | ···················· |
| S | 4260 hCoV-19/Spain/AS-IBV-98006641/2020\|EPI_ISL_654678\| | ···················· | ···················· | ···················· | ···················· | ···················· | ···················· |
| S | 4261 hCoV-19/Spain/AS-IBV-98006642/2020\|EPI_ISL_654679\| | ···················· | ···················· | ···················· | ···················· | ···················· | ···················· |
| S | 4262 hCoV-19/Spain/AS-IBV-98006655/2020\|EPI_ISL_654680\| | ···················· | ···················· | ···················· | ···················· | ···················· | ···················· |
| S | 4263 hCoV-19/Spain/AS-IBV-98006657/2020\|EPI_ISL_654681\| | ···················· | ···················· | ···················· | ···················· | ···················· | ···················· |
| S | 4264 hCoV-19/Spain/AS-IBV-98006591/2020\|EPI_ISL_654682\| | ···················· | ···················· | ···················· | ···················· | ···················· | ···················· |
| S | 4265 hCoV-19/Spain/AS-IBV-98006627/2020\|EPI_ISL_654683\| | ···················· | ···················· | ···················· | ···················· | ···················· | ···················· |
| S | 4266 hCoV-19/Spain/AS-IBV-98006637/2020\|EPI_ISL_654684\| | ···················· | ···················· | ···················· | ···················· | ···················· | ···················· |
| S | 4267 hCoV-19/USA/MN-MDH-2014/2020\|EPI_ISL_654695\|2020-0 | ···················· | ···················· | ···················· | ···················· | ···················· | ···················· |
| S | 4268 hCoV-19/USA/MN-MDH-2015/2020\|EPI_ISL_654696\|2020-0 | ···················· | ···················· | ···················· | ···················· | ···················· | ···················· |
| S | 4269 hCoV-19/USA/MN-MDH-2016/2020\|EPI_ISL_654697\|2020-0 | ···················· | ···················· | ···················· | ···················· | ···················· | ···················· |
| S | 4270 hCoV-19/USA/MN-MDH-2017/2020\|EPI_ISL_654698\|2020-0 | ···················· | ···················· | ···················· | ···················· | ···················· | ···················· |
| S | 4271 hCoV-19/USA/MN-MDH-2018/2020\|EPI_ISL_654699\|2020-1 | ···················· | ···················· | ···················· | ···················· | ···················· | ···················· |
| S | 4272 hCoV-19/USA/MN-MDH-2019/2020\|EPI_ISL_654700\|2020-1 | ···················· | ···················· | ···················· | ···················· | ···················· | ···················· |
| S | 4273 hCoV-19/USA/MN-MDH-2020/2020\|EPI_ISL_654701\|2020-1 | ···················· | ···················· | ···················· | ···················· | ···················· | ···················· |
| S | 4274 hCoV-19/USA/MN-MDH-2021/2020\|EPI_ISL_654702\|2020-1 | ···················· | ···················· | ···················· | ···················· | ···················· | ···················· |
| S | 4275 hCoV-19/USA/MN-MDH-2022/2020\|EPI_ISL_654703\|2020-1 | ···················· | ·A·················· | ···················· | ···················· | ···················· | ···················· |

| | | | | | | | |
|---|---|---|---|---|---|---|---|
| S | 4276 hCoV-19/USA/MN-MDH-2023/2020\|EPI_ISL_654704\|2020-1 | ···················· | ···················· | ···················· | ···················· | ···················· | ···················· |
| S | 4277 hCoV-19/USA/MN-MDH-2024/2020\|EPI_ISL_654705\|2020-1 | ···················· | ···················· | ···················· | ···················· | ···················· | ···················· |
| S | 4278 hCoV-19/USA/MN-MDH-2025/2020\|EPI_ISL_654706\|2020-1 | ···················· | ···················· | ···················· | ···················· | ···················· | ···················· |
| S | 4279 hCoV-19/USA/MN-MDH-2026/2020\|EPI_ISL_654707\|2020-1 | ···················· | ···················· | ···················· | ···················· | ···················· | ···················· |
| S | 4280 hCoV-19/USA/MN-MDH-2027/2020\|EPI_ISL_654708\|2020-1 | ···················· | ···················· | ···················· | ···················· | ···················· | ···················· |
| S | 4281 hCoV-19/USA/MN-MDH-2028/2020\|EPI_ISL_654709\|2020-1 | ···················· | ···················· | ···················· | ···················· | ···················· | ···················· |
| S | 4282 hCoV-19/USA/MN-MDH-2029/2020\|EPI_ISL_654710\|2020-1 | ···················· | ···················· | ···················· | ···················· | ···················· | ···················· |
| S | 4283 hCoV-19/USA/MN-MDH-2034/2020\|EPI_ISL_654715\|2020-1 | ···················· | ···················· | ···················· | ···················· | ···················· | ···················· |
| S | 4284 hCoV-19/USA/MN-MDH-2035/2020\|EPI_ISL_654716\|2020-1 | ···················· | ···················· | ···················· | ···················· | ···················· | ···················· |
| S | 4285 hCoV-19/USA/MN-MDH-2038/2020\|EPI_ISL_654719\|2020-1 | ···················· | ···················· | ···················· | ···················· | ···················· | ···················· |
| S | 4286 hCoV-19/USA/MN-MDH-2041/2020\|EPI_ISL_654722\|2020-1 | ···················· | ···················· | ···················· | ···················· | ···················· | ···················· |
| S | 4287 hCoV-19/USA/MN-MDH-2042/2020\|EPI_ISL_654723\|2020-1 | ···················· | ···················· | ···················· | ···················· | ···················· | ···················· |
| S | 4288 hCoV-19/USA/MN-MDH-2043/2020\|EPI_ISL_654724\|2020-1 | ···················· | ···················· | ···················· | ···················· | ···················· | ···················· |
| S | 4289 hCoV-19/USA/MN-MDH-2044/2020\|EPI_ISL_654725\|2020-1 | ···················· | ···················· | ···················· | ···················· | ···················· | ···················· |
| S | 4290 hCoV-19/USA/MN-MDH-2045/2020\|EPI_ISL_654726\|2020-1 | ···················· | ···················· | ···················· | ···················· | ···················· | ···················· |
| S | 4291 hCoV-19/USA/MN-MDH-2046/2020\|EPI_ISL_654727\|2020-1 | ···················· | ···················· | ···················· | ···················· | ······R············· | ···················· |
| S | 4292 hCoV-19/USA/MN-MDH-2047/2020\|EPI_ISL_654728\|2020-1 | ···················· | ···················· | ···················· | ···················· | ···················· | ···················· |
| S | 4293 hCoV-19/USA/MN-MDH-2048/2020\|EPI_ISL_654729\|2020-1 | ···················· | ···················· | ···················· | ···················· | ···················· | ·········N·········· |
| S | 4294 hCoV-19/England/ALDP-B5C93C/2020\|EPI_ISL_655965\|20 | ···················· | ···················· | ···················· | ···················· | ···················· | ···················· |
| S | 4295 hCoV-19/England/QEUH-B6C702/2020\|EPI_ISL_655967\|20 | ···················· | ···················· | ···················· | ···················· | ···················· | ···················· |
| S | 4296 hCoV-19/England/ALDP-B5C43B/2020\|EPI_ISL_655968\|20 | ···················· | ···················· | ···················· | ···················· | ···················· | ···················· |
| S | 4297 hCoV-19/England/ALDP-B5C9A5/2020\|EPI_ISL_655969\|20 | ···················· | ···················· | ···················· | ···················· | ···················· | ···················· |
| S | 4298 hCoV-19/England/ALDP-B5C486/2020\|EPI_ISL_655970\|20 | ···················· | ···················· | ···················· | ···················· | ···················· | ···················· |
| S | 4299 hCoV-19/England/QEUH-B6C6D8/2020\|EPI_ISL_655971\|20 | ···················· | ···················· | ···················· | ···················· | ···················· | ···················· |
| S | 4300 hCoV-19/England/ALDP-B5CF58/2020\|EPI_ISL_655972\|20 | ···················· | ·A·················· | ···················· | ···················· | ···················· | ···A················ |
| S | 4301 hCoV-19/England/QEUH-B65E8C/2020\|EPI_ISL_655973\|20 | ···················· | ···················· | ···················· | ···················· | ···················· | ···················· |
| S | 4302 hCoV-19/England/ALDP-B5CED3/2020\|EPI_ISL_655974\|20 | ···················· | ···················· | ···················· | ···················· | ···················· | ···················· |
| S | 4303 hCoV-19/England/QEUH-B6C76C/2020\|EPI_ISL_655975\|20 | ············T······· | ···················· | ···················· | ···················· | ···················· | ···················· |
| S | 4304 hCoV-19/England/ALDP-B5CB45/2020\|EPI_ISL_655976\|20 | ···················· | ···················· | ···················· | ···················· | ···················· | ···················· |
| S | 4305 hCoV-19/England/ALDP-B5CC42/2020\|EPI_ISL_655977\|20 | ···················· | ···················· | ···················· | ···················· | ···················· | ···················· |
| S | 4306 hCoV-19/England/ALDP-B46B5D/2020\|EPI_ISL_655978\|20 | ···················· | ···················· | ···················· | ···················· | ···················· | ···················· |
| S | 4307 hCoV-19/England/ALDP-B46BD5/2020\|EPI_ISL_655979\|20 | ···················· | ···················· | ···················· | ···················· | ···················· | ···················· |
| S | 4308 hCoV-19/England/ALDP-B45B6D/2020\|EPI_ISL_655980\|20 | ···················· | ···················· | ···················· | ···················· | ···················· | ···················· |
| S | 4309 hCoV-19/England/ALDP-B468CF/2020\|EPI_ISL_655981\|20 | ···················· | ···················· | ···················· | ···················· | ···················· | ···················· |
| S | 4310 hCoV-19/England/ALDP-B4664D/2020\|EPI_ISL_655982\|20 | ···················· | ···················· | ···················· | ···················· | ···················· | ···················· |
| S | 4311 hCoV-19/England/ALDP-B46971/2020\|EPI_ISL_655984\|20 | ···················· | ···················· | ···················· | ···················· | ···················· | ···················· |
| S | 4312 hCoV-19/England/ALDP-B462C1/2020\|EPI_ISL_655985\|20 | ···················· | ···················· | ···················· | ···················· | ···················· | ···················· |
| S | 4313 hCoV-19/England/ALDP-B45F9E/2020\|EPI_ISL_655986\|20 | ···················· | ···················· | ···················· | ···················· | ···················· | ···················· |
| S | 4314 hCoV-19/England/ALDP-B46504/2020\|EPI_ISL_655987\|20 | ···················· | ···················· | ···················· | ···················· | ···················· | ···················· |
| S | 4315 hCoV-19/England/ALDP-B4616A/2020\|EPI_ISL_655988\|20 | ···················· | ···················· | ···················· | ···················· | ···················· | ···················· |
| S | 4316 hCoV-19/England/ALDP-B46D66/2020\|EPI_ISL_655989\|20 | ···················· | ···················· | ···················· | ···················· | ···················· | ···················· |
| S | 4317 hCoV-19/England/ALDP-B45BA9/2020\|EPI_ISL_655990\|20 | ···················· | ···················· | ···················· | ···················· | ···················· | ···················· |
| S | 4318 hCoV-19/England/ALDP-B45954/2020\|EPI_ISL_655991\|20 | ···················· | ···················· | ···················· | ···················· | ···················· | ···················· |
| S | 4319 hCoV-19/England/ALDP-B469BD/2020\|EPI_ISL_655993\|20 | ···················· | ···················· | ···················· | ···················· | ···················· | ···················· |
| S | 4320 hCoV-19/England/ALDP-B46A32/2020\|EPI_ISL_655994\|20 | ···················· | ···················· | ···················· | ···················· | ···················· | ···················· |
| S | 4321 hCoV-19/England/ALDP-B46944/2020\|EPI_ISL_655996\|20 | ···················· | ···················· | ···················· | ···················· | ···················· | ···················· |
| S | 4322 hCoV-19/England/ALDP-B45EDD/2020\|EPI_ISL_655997\|20 | ···················· | ···················· | ···················· | ···················· | ···················· | ···················· |
| S | 4323 hCoV-19/England/ALDP-B46531/2020\|EPI_ISL_655998\|20 | ···················· | ···················· | ···················· | ···················· | ···················· | ···················· |
| S | 4324 hCoV-19/England/ALDP-B4655F/2020\|EPI_ISL_655999\|20 | ···················· | ···················· | ···················· | ···················· | ···················· | ···················· |
| S | 4325 hCoV-19/England/MILK-B561A3/2020\|EPI_ISL_658280\|20 | ···················· | ···················· | ···················· | ···················· | ···················· | ···················· |
| S | 4326 hCoV-19/England/MILK-B5A5D0/2020\|EPI_ISL_658281\|20 | ···················· | ···················· | ···················· | ···················· | ···················· | ···················· |
| S | 4327 hCoV-19/England/MILK-B55EE9/2020\|EPI_ISL_658282\|20 | ···················· | ···················· | ···················· | ···················· | ···················· | ···················· |
| S | 4328 hCoV-19/England/MILK-B55C49/2020\|EPI_ISL_658283\|20 | ···················· | ···················· | ···················· | ···················· | ···················· | ···················· |
| S | 4329 hCoV-19/England/MILK-B55E07/2020\|EPI_ISL_658284\|20 | ···················· | ···················· | ···················· | ···················· | ···················· | ···················· |
| S | 4330 hCoV-19/England/CAMC-B5183A/2020\|EPI_ISL_658285\|20 | ···················· | ···················· | ···················· | ···················· | ···················· | ···················· |
| S | 4331 hCoV-19/England/CAMC-B54794/2020\|EPI_ISL_658286\|20 | ···················· | ···················· | ···················· | ···················· | ······T············· | ···················· |
| S | 4332 hCoV-19/England/CAMC-B517D3/2020\|EPI_ISL_658287\|20 | ···················· | ···················· | ···················· | ···················· | ···················· | ···················· |
| S | 4333 hCoV-19/England/MILK-B5612B/2020\|EPI_ISL_658288\|20 | ···················· | ······R············· | ···················· | ···················· | ···················· | ···················· |
| S | 4334 hCoV-19/England/MILK-B5A5A3/2020\|EPI_ISL_658290\|20 | ···················· | ···················· | ···················· | ···················· | ···················· | ···················· |
| S | 4335 hCoV-19/England/MILK-B5A716/2020\|EPI_ISL_658291\|20 | ···················· | ···················· | ···················· | ···················· | ······T············· | ···················· |
| S | 4336 hCoV-19/England/MILK-B55CB2/2020\|EPI_ISL_658293\|20 | ···················· | ···················· | ···················· | ···················· | ···················· | ···················· |
| S | 4337 hCoV-19/England/MILK-B5A549/2020\|EPI_ISL_658294\|20 | ···················· | ···················· | ···················· | ···················· | ···················· | ···················· |
| S | 4338 hCoV-19/England/MILK-B5A619/2020\|EPI_ISL_658295\|20 | ···················· | ···················· | ···················· | ···················· | ···················· | ···················· |
| S | 4339 hCoV-19/England/MILK-B55F04/2020\|EPI_ISL_658296\|20 | ···················· | ···················· | ···················· | ···················· | ···················· | ···················· |
| S | 4340 hCoV-19/England/MILK-B55FE6/2020\|EPI_ISL_658297\|20 | ···················· | ·A·······A·········· | ···················· | ···················· | ···················· | ···················· |
| S | 4341 hCoV-19/England/MILK-B5A5EF/2020\|EPI_ISL_658298\|20 | ···················· | ···················· | ···················· | ···················· | ···················· | ···················· |
| S | 4342 hCoV-19/England/CAMC-B5175B/2020\|EPI_ISL_658299\|20 | ···················· | ···················· | ···················· | ···················· | ···················· | ···················· |
| S | 4343 hCoV-19/England/CAMC-B351F5/2020\|EPI_ISL_659511\|20 | ···················· | ···················· | ···················· | ···················· | ···················· | ···················· |
| S | 4344 hCoV-19/England/CAMC-B34958/2020\|EPI_ISL_659512\|20 | ···················· | ···················· | ···················· | ···················· | ···················· | ···················· |
| S | 4345 hCoV-19/England/CAMC-B353C2/2020\|EPI_ISL_659513\|20 | ···················· | ···················· | ···················· | ···················· | ······T············· | ···················· |
| S | 4346 hCoV-19/England/CAMC-B355AE/2020\|EPI_ISL_659515\|20 | ···················· | ·A·················· | ···················· | ···················· | ···················· | ···A················ |
| S | 4347 hCoV-19/England/CAMC-B344FD/2020\|EPI_ISL_659516\|20 | ···················· | ···················· | ···················· | ···················· | ···················· | ···················· |
| S | 4348 hCoV-19/England/CAMC-B34DC5/2020\|EPI_ISL_659517\|20 | ···················· | ···················· | ···················· | ···················· | ···················· | ···················· |
| S | 4349 hCoV-19/Czech Republic/NRL-9963/2020\|EPI_ISL_66059 | ···················· | ···················· | ···················· | ···················· | ···················· | ···················· |
| S | 4350 hCoV-19/Czech Republic/NRL-9964/2020\|EPI_ISL_66059 | ···················· | ···················· | ···················· | ···················· | ···················· | ···················· |
| S | 4351 hCoV-19/Czech Republic/NRL-10066/2020\|EPI_ISL_6605 | ···················· | ···················· | ···················· | ···················· | ···················· | ···················· |
| S | 4352 hCoV-19/Czech Republic/NRL-9925/2020\|EPI_ISL_66059 | ···················· | ···················· | ···················· | ···················· | ···················· | ···················· |
| S | 4353 hCoV-19/England/ALDP-B48F7D/2020\|EPI_ISL_649200\|20 | ···················· | ···················· | ···················· | ···················· | ···················· | ···················· |
| S | 4354 hCoV-19/England/ALDP-B48FD7/2020\|EPI_ISL_649201\|20 | ···················· | ···················· | ···················· | ···················· | ···················· | ···················· |
| S | 4355 hCoV-19/England/ALDP-B48283/2020\|EPI_ISL_649202\|20 | ···················· | ···················· | ···················· | ···················· | ···················· | ···················· |

| | | | | | | | | |
|---|---|---|---|---|---|---|---|---|
| S | 4356 | hCoV-19/England/ALDP-B47EDB/2020\|EPI_ISL_649203\|20 | ···················· | ···················· | ····················· | ···················· | ······T············· | ···················· |
| S | 4357 | hCoV-19/England/ALDP-B48854/2020\|EPI_ISL_649204\|20 | ···················· | ·A··G··············· | ····················· | ···················· | ···················· | ···················· |
| S | 4358 | hCoV-19/England/MILK-B50561/2020\|EPI_ISL_649205\|20 | ···················· | ···················· | ····················· | ···················· | ···················· | ···················· |
| S | 4359 | hCoV-19/England/ALDP-B493BC/2020\|EPI_ISL_649206\|20 | ···················· | ···················· | ····················· | ···················· | ···················· | ···················· |
| S | 4360 | hCoV-19/England/ALDP-B48E43/2020\|EPI_ISL_649207\|20 | ···················· | ···················· | ····················· | ···················· | ···················· | ···················· |
| S | 4361 | hCoV-19/England/ALDP-B486C3/2020\|EPI_ISL_649208\|20 | ···················· | ···················· | ····················· | ···················· | ···················· | ···················· |
| S | 4362 | hCoV-19/England/ALDP-B48863/2020\|EPI_ISL_649209\|20 | ···················· | ···················· | ····················· | ···················· | ···················· | ···················· |
| S | 4363 | hCoV-19/England/ALDP-B47D83/2020\|EPI_ISL_649230\|20 | ···················· | ···················· | ····················· | ···················· | ···················· | ···················· |
| S | 4364 | hCoV-19/England/ALDP-B482DE/2020\|EPI_ISL_649231\|20 | ···················· | ···················· | ····················· | ···················· | ···················· | ···················· |
| S | 4365 | hCoV-19/England/ALDP-B480D4/2020\|EPI_ISL_649232\|20 | ···················· | ···················· | ····················· | ···················· | ···················· | ···················· |
| S | 4366 | hCoV-19/England/ALDP-B48DBF/2020\|EPI_ISL_649233\|20 | ···················· | ···················· | ····················· | ···················· | ···················· | ···················· |
| S | 4367 | hCoV-19/England/ALDP-B50A17/2020\|EPI_ISL_649234\|20 | ···················· | ···················· | ····················· | ···················· | ···················· | ···················· |
| S | 4368 | hCoV-19/England/ALDP-B48380/2020\|EPI_ISL_649235\|20 | ···················· | ···················· | ····················· | ···················· | ···················· | ···················· |
| S | 4369 | hCoV-19/England/ALDP-B4905B/2020\|EPI_ISL_649237\|20 | ···················· | ·········T·········· | ····················· | ···················· | ···················· | ···················· |
| S | 4370 | hCoV-19/England/ALDP-B49158/2020\|EPI_ISL_649238\|20 | ···················· | ···················· | ····················· | ···················· | ···················· | ···················· |
| S | 4371 | hCoV-19/England/ALDP-B4860F/2020\|EPI_ISL_649239\|20 | ···················· | ···················· | ····················· | ···················· | ···················· | ···A················ |
| S | 4372 | hCoV-19/England/ALDP-B485D5/2020\|EPI_ISL_649250\|20 | ···················· | ···················· | ····················· | ···················· | ···················· | ···················· |
| S | 4373 | hCoV-19/England/ALDP-B47F14/2020\|EPI_ISL_649251\|20 | ···················· | ···················· | ····················· | ···················· | ···················· | ···················· |
| S | 4374 | hCoV-19/England/ALDP-B50CE4/2020\|EPI_ISL_649252\|20 | ···················· | ···················· | ····················· | ···················· | ···················· | ···················· |
| S | 4375 | hCoV-19/England/ALDP-B47DB0/2020\|EPI_ISL_649253\|20 | ···················· | ·········T·········· | ····················· | ···················· | ···················· | ···················· |
| S | 4376 | hCoV-19/England/ALDP-B483F9/2020\|EPI_ISL_649254\|20 | ···················· | ···················· | ····················· | ···················· | ···················· | ···················· |
| S | 4377 | hCoV-19/England/MILK-B504FB/2020\|EPI_ISL_649255\|20 | ···················· | ···················· | ····················· | ···················· | ···················· | ···················· |
| S | 4378 | hCoV-19/England/ALDP-B490B5/2020\|EPI_ISL_649256\|20 | ···················· | ···················· | ····················· | ···················· | ···················· | ···················· |
| S | 4379 | hCoV-19/England/ALDP-B4FC51/2020\|EPI_ISL_649257\|20 | ···················· | ···················· | ····················· | ···················· | ···················· | ···················· |
| S | 4380 | hCoV-19/England/ALDP-B48696/2020\|EPI_ISL_649259\|20 | ···················· | ···················· | ····················· | ···················· | ···················· | ···················· |
| S | 4381 | hCoV-19/England/ALDP-B47DFC/2020\|EPI_ISL_649260\|20 | ···················· | ···················· | ····················· | ···················· | ···················· | ···················· |
| S | 4382 | hCoV-19/England/ALDP-B507F2/2020\|EPI_ISL_649261\|20 | ···················· | ···················· | ····················· | ···················· | ···················· | ···················· |
| S | 4383 | hCoV-19/England/ALDP-B47BB6/2020\|EPI_ISL_649263\|20 | ···················· | ···················· | ····················· | ···················· | ···················· | ···················· |
| S | 4384 | hCoV-19/England/ALDP-B4861E/2020\|EPI_ISL_649264\|20 | ···················· | ···················· | ····················· | ···················· | ···················· | ···················· |
| S | 4385 | hCoV-19/England/ALDP-B49325/2020\|EPI_ISL_649265\|20 | ···················· | ···················· | ····················· | ···················· | ···················· | ···················· |
| S | 4386 | hCoV-19/England/ALDP-B49307/2020\|EPI_ISL_649266\|20 | ···················· | ·A·················· | ····················· | ···················· | ···················· | ···A················ |
| S | 4387 | hCoV-19/England/ALDP-B481D1/2020\|EPI_ISL_649267\|20 | ···················· | ···················· | ····················· | ···················· | ···················· | ···················· |
| S | 4388 | hCoV-19/England/ALDP-B484E7/2020\|EPI_ISL_649268\|20 | ···················· | ···················· | ····················· | ···················· | ···················· | ···················· |
| S | 4389 | hCoV-19/England/ALDP-B48308/2020\|EPI_ISL_649269\|20 | ···················· | ···················· | ····················· | ···················· | ···················· | ···················· |
| S | 4390 | hCoV-19/England/ALDP-B482CF/2020\|EPI_ISL_649270\|20 | ···················· | ···················· | ····················· | ···················· | ···················· | ···················· |
| S | 4391 | hCoV-19/England/ALDP-B493F8/2020\|EPI_ISL_649271\|20 | ···················· | ···················· | ····················· | ···················· | ···················· | ···················· |
| S | 4392 | hCoV-19/England/ALDP-B48F31/2020\|EPI_ISL_649273\|20 | ···················· | ···················· | ····················· | ···················· | ···················· | ···················· |
| S | 4393 | hCoV-19/England/ALDP-B47D38/2020\|EPI_ISL_649274\|20 | ···················· | ···················· | ····················· | ···················· | ···················· | ···················· |
| S | 4394 | hCoV-19/England/ALDP-B48EAD/2020\|EPI_ISL_649275\|20 | ···················· | ···················· | ····················· | ···················· | ···················· | ···················· |
| S | 4395 | hCoV-19/England/ALDP-B49097/2020\|EPI_ISL_649276\|20 | ···················· | ···················· | ····················· | ···················· | ···················· | ···················· |
| S | 4396 | hCoV-19/England/ALDP-B480A7/2020\|EPI_ISL_649277\|20 | ···················· | ···················· | ····················· | ···················· | ···················· | ···················· |
| S | 4397 | hCoV-19/England/ALDP-B4871B/2020\|EPI_ISL_649278\|20 | ···················· | ···················· | ····················· | ···················· | ···················· | ···················· |
| S | 4398 | hCoV-19/England/ALDP-B491A3/2020\|EPI_ISL_649279\|20 | ···················· | ···················· | ····················· | ···················· | ···················· | ···················· |
| S | 4399 | hCoV-19/England/ALDP-B47F6F/2020\|EPI_ISL_649280\|20 | ···················· | ···················· | ····················· | ···················· | ···················· | ···················· |
| S | 4400 | hCoV-19/England/ALDP-B48229/2020\|EPI_ISL_649281\|20 | ···················· | ···················· | ····················· | ···················· | ···················· | ···················· |
| S | 4401 | hCoV-19/England/ALDP-B48326/2020\|EPI_ISL_649282\|20 | ···················· | ···················· | ····················· | ···················· | ···················· | ···················· |
| S | 4402 | hCoV-19/England/MILK-B50552/2020\|EPI_ISL_649283\|20 | ···················· | ···················· | ····················· | ···················· | ···················· | ···················· |
| S | 4403 | hCoV-19/England/ALDP-B4904C/2020\|EPI_ISL_649284\|20 | ···················· | ·········T·········· | ····················· | ···················· | ···················· | ···················· |
| S | 4404 | hCoV-19/England/ALDP-B49167/2020\|EPI_ISL_649285\|20 | ···················· | ···················· | ····················· | ···················· | ···················· | ···················· |
| S | 4405 | hCoV-19/England/ALDP-B4902E/2020\|EPI_ISL_649287\|20 | ···················· | ···················· | ····················· | ···················· | ···················· | ···················· |
| S | 4406 | hCoV-19/England/ALDP-B4855D/2020\|EPI_ISL_649289\|20 | ···················· | ···················· | ····················· | ···················· | ···················· | ···················· |
| S | 4407 | hCoV-19/England/BIRM-675DB/2020\|EPI_ISL_651519\|202 | ···················· | ···················· | ····················· | ···················· | ···················· | ···················· |
| S | 4408 | hCoV-19/Scotland/EDB8208/2020\|EPI_ISL_651520\|2020- | ···················· | ···················· | ····················· | ···················· | ···················· | ···················· |
| S | 4409 | hCoV-19/England/BIRM-640A0/2020\|EPI_ISL_651590\|202 | ···················· | ···················· | ····················· | ···················· | ···················· | ···················· |
| S | 4410 | hCoV-19/England/BIRM-640CE/2020\|EPI_ISL_651591\|202 | ···················· | ···················· | ····················· | ···················· | ···················· | ···················· |
| S | 4411 | hCoV-19/England/BIRM-640DD/2020\|EPI_ISL_651592\|202 | ···················· | ···················· | ····················· | ···················· | ···················· | ···················· |
| S | 4412 | hCoV-19/England/BIRM-640FB/2020\|EPI_ISL_651593\|202 | ···················· | ···················· | ····················· | ···················· | ···················· | ···················· |
| S | 4413 | hCoV-19/England/BIRM-64107/2020\|EPI_ISL_651594\|202 | ···················· | ···················· | ····················· | ···················· | ···················· | ···················· |
| S | 4414 | hCoV-19/England/BIRM-64116/2020\|EPI_ISL_651595\|202 | ···················· | ···················· | ····················· | ···················· | ···················· | ···················· |
| S | 4415 | hCoV-19/England/BIRM-64134/2020\|EPI_ISL_651596\|202 | ···················· | ···················· | ····················· | ···················· | ···················· | ···················· |
| S | 4419 | hCoV-19/Scotland/EDB9460/2020\|EPI_ISL_652060\|2020- | ···················· | ···················· | ····················· | ···················· | ···················· | ···················· |
| S | 4420 | hCoV-19/Scotland/EDB9479/2020\|EPI_ISL_652061\|2020- | ···················· | ···················· | ····················· | ···················· | ···················· | ···················· |
| S | 4421 | hCoV-19/Scotland/EDB9489/2020\|EPI_ISL_652062\|2020- | ···················· | ···················· | ····················· | ···················· | ···················· | ···················· |
| S | 4422 | hCoV-19/Scotland/EDB9494/2020\|EPI_ISL_652063\|2020- | ···················· | ···················· | ····················· | ···················· | ···················· | ···················· |
| S | 4423 | hCoV-19/Scotland/EDB9506/2020\|EPI_ISL_652064\|2020- | ···················· | ···················· | ····················· | ···················· | ···················· | ···················· |
| S | 4424 | hCoV-19/Scotland/EDB9507/2020\|EPI_ISL_652065\|2020- | ···················· | ···················· | ····················· | ···················· | ···················· | ···················· |
| S | 4425 | hCoV-19/Scotland/EDB9508/2020\|EPI_ISL_652066\|2020- | ···················· | ···················· | ····················· | ···················· | ···················· | ···················· |
| S | 4426 | hCoV-19/England/HECH-259A7B0/2020\|EPI_ISL_652067\|2 | ···················· | ···················· | ····················· | ···················· | ···················· | ···················· |
| S | 4427 | hCoV-19/England/HECH-259A905/2020\|EPI_ISL_652068\|2 | ···················· | ···················· | ····················· | ···················· | ···················· | ···················· |
| S | 4428 | hCoV-19/England/HECH-259A923/2020\|EPI_ISL_652069\|2 | ···················· | ···················· | ····················· | ···················· | ···················· | ···················· |
| S | 4429 | hCoV-19/England/HECH-259A950/2020\|EPI_ISL_652070\|2 | ···················· | ···················· | ····················· | ···················· | ···················· | ···················· |
| S | 4430 | hCoV-19/England/HECH-259AAA8/2020\|EPI_ISL_652071\|2 | ···················· | ···················· | ····················· | ···················· | ···················· | ···················· |
| S | 4431 | hCoV-19/England/HECH-259AAC6/2020\|EPI_ISL_652072\|2 | ···················· | ···················· | ····················· | ···················· | ···················· | ···················· |
| S | 4432 | hCoV-19/England/LIVE-DB38E9/2020\|EPI_ISL_652073\|20 | ···················· | ···················· | ····················· | ···················· | ···················· | ···················· |
| S | 4433 | hCoV-19/England/LIVE-DB3BB3/2020\|EPI_ISL_652074\|20 | ···················· | ···················· | ····················· | ···················· | ···················· | ···················· |
| S | 4434 | hCoV-19/England/LIVE-DB3C74/2020\|EPI_ISL_652075\|20 | ···················· | ···················· | ····················· | ···················· | ···················· | ···················· |
| S | 4435 | hCoV-19/England/LIVE-DB3D17/2020\|EPI_ISL_652076\|20 | ···················· | ·········T·········· | ····················· | ···················· | ···················· | ···················· |
| S | 4436 | hCoV-19/England/LIVE-DB3DAE/2020\|EPI_ISL_652077\|20 | ···················· | ···················· | ····················· | ···················· | ···················· | ···················· |
| S | 4437 | hCoV-19/England/LIVE-DB40B3/2020\|EPI_ISL_652078\|20 | ···················· | ···················· | ····················· | ···················· | ···················· | ···················· |
| S | 4438 | hCoV-19/England/LIVE-DB40C2/2020\|EPI_ISL_652079\|20 | ···················· | ···················· | ····················· | ···················· | ···················· | ···················· |

| | | | | | | | |
|---|---|---|---|---|---|---|---|
| S | 4439 hCoV-19/England/LIVE-DB40E0/2020\|EPI_ISL_652080\|20 | ···················· | ···················· | ···················· | ···················· | ···················· | ···················· |
| S | 4440 hCoV-19/England/LIVE-DB41B0/2020\|EPI_ISL_652081\|20 | ···················· | ···················· | ···················· | ···················· | ···················· | ···················· |
| S | 4441 hCoV-19/England/LIVE-DB41CF/2020\|EPI_ISL_652082\|20 | ········Y··········· | ·········T·········· | ···················· | ···················· | ···················· | ···················· |
| S | 4442 hCoV-19/England/LIVE-DB41DE/2020\|EPI_ISL_652083\|20 | ···················· | ·········T·········· | ···················· | ···················· | ···················· | ···················· |
| S | 4443 hCoV-19/England/LIVE-DB41FC/2020\|EPI_ISL_652084\|20 | ···················· | ···················· | ···················· | ···················· | ···················· | ···················· |
| S | 4444 hCoV-19/England/LIVE-DB436F/2020\|EPI_ISL_652085\|20 | ···················· | ···················· | ···················· | ···················· | ···················· | ···················· |
| S | 4445 hCoV-19/England/LIVE-DB43C9/2020\|EPI_ISL_652086\|20 | ···················· | ···················· | ···················· | ···················· | ···················· | ···················· |
| S | 4446 hCoV-19/England/LIVE-DB4B49/2020\|EPI_ISL_652087\|20 | ···················· | ···················· | ···················· | ···················· | ···················· | ···················· |
| S | 4447 hCoV-19/England/LIVE-DB4BFE/2020\|EPI_ISL_652088\|20 | ···················· | ···················· | ···················· | ···················· | ···················· | ···················· |
| S | 4448 hCoV-19/England/LIVE-DB601B/2020\|EPI_ISL_652089\|20 | ···················· | ···················· | ···················· | ···················· | ···················· | ···················· |
| S | 4449 hCoV-19/England/LIVE-DB6039/2020\|EPI_ISL_652090\|20 | ···················· | ···················· | ···················· | ···················· | ···················· | ···················· |
| S | 4450 hCoV-19/England/LIVE-DB6057/2020\|EPI_ISL_652091\|20 | ···················· | ···················· | ···················· | ···················· | ···················· | ···················· |
| S | 4451 hCoV-19/England/LIVE-DB6093/2020\|EPI_ISL_652092\|20 | ···················· | ···················· | ···················· | ···················· | ···················· | ···················· |
| S | 4452 hCoV-19/England/LIVE-DB60C0/2020\|EPI_ISL_652093\|20 | ···················· | ···················· | ···················· | ···················· | ···················· | ···················· |
| S | 4453 hCoV-19/England/LIVE-DB61DC/2020\|EPI_ISL_652094\|20 | ···················· | ···················· | ···················· | ···················· | ···················· | ···················· |
| S | 4454 hCoV-19/England/LIVE-DB6242/2020\|EPI_ISL_652095\|20 | ···················· | ···················· | ···················· | ···················· | ···················· | ···················· |
| S | 4455 hCoV-19/England/LIVE-DB635E/2020\|EPI_ISL_652096\|20 | ···················· | ···················· | ···················· | ···················· | ···················· | ···················· |
| S | 4456 hCoV-19/England/LIVE-DB644C/2020\|EPI_ISL_652097\|20 | ···················· | ···················· | ···················· | ···················· | ···················· | ···················· |
| S | 4457 hCoV-19/England/LIVE-DB645B/2020\|EPI_ISL_652098\|20 | ···················· | ···················· | ···················· | ···················· | ···················· | ···················· |
| S | 4458 hCoV-19/England/LIVE-DB646A/2020\|EPI_ISL_652099\|20 | ···················· | ···················· | ···················· | ···················· | ···················· | ···················· |
| S | 4459 hCoV-19/Scotland/QEUH-AAB5FF/2020\|EPI_ISL_652800\|2 | ···················· | ···················· | ···················· | ···················· | ···················· | ···················· |
| S | 4460 hCoV-19/Scotland/QEUH-AAB61A/2020\|EPI_ISL_652801\|2 | ···················· | ···················· | ···················· | ···················· | ···················· | ···················· |
| S | 4461 hCoV-19/Scotland/QEUH-AAB629/2020\|EPI_ISL_652802\|2 | ···················· | ···················· | ···················· | ···················· | ···················· | ···················· |
| S | 4462 hCoV-19/England/QEUH-AAB674/2020\|EPI_ISL_652803\|20 | ···················· | ·············C······ | ···················· | ···················· | ···················· | ···················· |
| S | 4463 hCoV-19/England/QEUH-AAB683/2020\|EPI_ISL_652804\|20 | ···················· | ···················· | ···················· | ···················· | ···················· | ···················· |
| S | 4464 hCoV-19/England/QEUH-AAB6A1/2020\|EPI_ISL_652805\|20 | ···················· | ···················· | ···················· | ···················· | ···················· | ···················· |
| S | 4465 hCoV-19/England/QEUH-AAB6B0/2020\|EPI_ISL_652806\|20 | ···················· | ···················· | ···················· | ···················· | ···················· | ···················· |
| S | 4466 hCoV-19/England/QEUH-AAB6CF/2020\|EPI_ISL_652807\|20 | ···················· | ·A·················· | ···················· | ···················· | ···················· | ···················· |
| S | 4467 hCoV-19/Wales/QEUH-AAB726/2020\|EPI_ISL_652808\|2020 | ···················· | ···················· | ···················· | ···················· | ···················· | ···················· |
| S | 4468 hCoV-19/England/QEUH-AAB744/2020\|EPI_ISL_652809\|20 | ···················· | ···················· | ···················· | ···················· | ···················· | ···················· |
| S | 4469 hCoV-19/England/QEUH-AAB753/2020\|EPI_ISL_652810\|20 | ···················· | ···················· | ···················· | ···················· | ···················· | ···················· |
| S | 4470 hCoV-19/England/QEUH-AAB771/2020\|EPI_ISL_652811\|20 | ···················· | ···················· | ···················· | ···················· | ···················· | ···················· |
| S | 4471 hCoV-19/England/QEUH-AAB780/2020\|EPI_ISL_652812\|20 | ···················· | ···················· | ···················· | ···················· | ···················· | ···················· |
| S | 4472 hCoV-19/England/QEUH-AAB7AE/2020\|EPI_ISL_652813\|20 | ···················· | ···················· | ···················· | ···················· | ···················· | ···················· |
| S | 4473 hCoV-19/England/QEUH-AAB7DB/2020\|EPI_ISL_652814\|20 | ···················· | ···················· | ···················· | ···················· | ···················· | ···················· |
| S | 4474 hCoV-19/England/QEUH-AAB814/2020\|EPI_ISL_652815\|20 | ···················· | ···················· | ···················· | ···················· | ···················· | ···················· |
| S | 4475 hCoV-19/England/QEUH-AAB823/2020\|EPI_ISL_652816\|20 | ···················· | ···················· | ···················· | ···················· | ···················· | ···················· |
| S | 4476 hCoV-19/England/QEUH-AAB841/2020\|EPI_ISL_652817\|20 | ···················· | ···················· | ···················· | ···················· | ···················· | ···················· |
| S | 4477 hCoV-19/England/QEUH-AABB66/2020\|EPI_ISL_652818\|20 | ···················· | ···················· | ···················· | ···················· | ···················· | ···················· |
| S | 4478 hCoV-19/England/QEUH-AABC90/2020\|EPI_ISL_652819\|20 | ···················· | ···················· | ···················· | ···················· | ···················· | ···················· |
| S | 4479 hCoV-19/England/QEUH-AABCBE/2020\|EPI_ISL_652820\|20 | ···················· | ···················· | ···················· | ···················· | ···················· | ···················· |
| S | 4480 hCoV-19/England/QEUH-AABCCD/2020\|EPI_ISL_652821\|20 | ···················· | ···················· | ···················· | ···················· | ···················· | ···················· |
| S | 4481 hCoV-19/England/QEUH-AABCDC/2020\|EPI_ISL_652822\|20 | ···················· | ···················· | ···················· | ···················· | ···················· | ···················· |
| S | 4482 hCoV-19/England/QEUH-AABCEB/2020\|EPI_ISL_652823\|20 | ···················· | ···················· | ···················· | ···················· | ···················· | ···················· |
| S | 4483 hCoV-19/England/QEUH-AABCFA/2020\|EPI_ISL_652824\|20 | ···················· | ···················· | ···················· | ···················· | ···················· | ···················· |
| S | 4484 hCoV-19/England/QEUH-AABD15/2020\|EPI_ISL_652825\|20 | ···················· | ···················· | ···················· | ···················· | ···················· | ···················· |
| S | 4485 hCoV-19/England/QEUH-AABD24/2020\|EPI_ISL_652826\|20 | ···················· | ···················· | ···················· | ···················· | ···················· | ···················· |
| S | 4486 hCoV-19/England/QEUH-AABD33/2020\|EPI_ISL_652827\|20 | ···················· | ···················· | ···················· | ···················· | ···················· | ···················· |
| S | 4487 hCoV-19/England/QEUH-AABD51/2020\|EPI_ISL_652828\|20 | ···················· | ···················· | ···················· | ···················· | ···················· | ···················· |
| S | 4488 hCoV-19/England/QEUH-AABD60/2020\|EPI_ISL_652829\|20 | ···················· | ···················· | ···················· | ···················· | ···················· | ···················· |
| S | 4489 hCoV-19/England/QEUH-AABD8E/2020\|EPI_ISL_652830\|20 | ···················· | ···················· | ···················· | ···················· | ···················· | ···················· |
| S | 4490 hCoV-19/England/QEUH-AABD9D/2020\|EPI_ISL_652831\|20 | ···················· | ···················· | ···················· | ···················· | ···················· | ···················· |
| S | 4491 hCoV-19/England/QEUH-AABDBB/2020\|EPI_ISL_652832\|20 | ···················· | ···················· | ···················· | ···················· | ···················· | ···················· |
| S | 4492 hCoV-19/England/QEUH-AABDCA/2020\|EPI_ISL_652833\|20 | ···················· | ···················· | ···················· | ···················· | ···················· | ···················· |
| S | 4493 hCoV-19/England/QEUH-AABDE8/2020\|EPI_ISL_652834\|20 | ···················· | ···················· | ···················· | ···················· | ···················· | ···················· |
| S | 4494 hCoV-19/Scotland/QEUH-AABF2E/2020\|EPI_ISL_652835\|2 | ···················· | ···················· | ···················· | ···················· | ···················· | ···················· |
| S | 4495 hCoV-19/Scotland/QEUH-AABF5B/2020\|EPI_ISL_652836\|2 | ···················· | ···················· | ···················· | ···················· | ···················· | ···················· |
| S | 4496 hCoV-19/Scotland/QEUH-AABF6A/2020\|EPI_ISL_652837\|2 | ···················· | ···················· | ···················· | ···················· | ···················· | ···················· |
| S | 4497 hCoV-19/Scotland/QEUH-AABF88/2020\|EPI_ISL_652838\|2 | ···················· | ···················· | ···················· | ···················· | ······T············· | ···················· |
| S | 4498 hCoV-19/Scotland/QEUH-AABFB5/2020\|EPI_ISL_652839\|2 | ···················· | ···················· | ···················· | ···················· | ···················· | ···················· |
| S | 4499 hCoV-19/Scotland/QEUH-AABFC4/2020\|EPI_ISL_652840\|2 | ···················· | ···················· | ···················· | ···················· | ···················· | ···················· |
| S | 4500 hCoV-19/Scotland/QEUH-AABFD3/2020\|EPI_ISL_652841\|2 | ···················· | ···················· | ···················· | ···················· | ···················· | ···················· |
| S | 4501 hCoV-19/Scotland/QEUH-AABFE2/2020\|EPI_ISL_652842\|2 | ···················· | ···················· | ···················· | ···················· | ···················· | ···················· |
| S | 4502 hCoV-19/England/QEUH-AAC02A/2020\|EPI_ISL_652843\|20 | ···················· | ···················· | ···················· | ···················· | ···················· | ···················· |
| S | 4503 hCoV-19/England/QEUH-AAC066/2020\|EPI_ISL_652844\|20 | ···················· | ···················· | ···················· | ···················· | ···················· | ···················· |
| S | 4504 hCoV-19/England/QEUH-AAC136/2020\|EPI_ISL_652845\|20 | ···················· | ···················· | ···················· | ···················· | ···················· | ···················· |
| S | 4505 hCoV-19/England/QEUH-AAC1BE/2020\|EPI_ISL_652846\|20 | ···················· | ···················· | ···················· | ···················· | ···················· | ···················· |
| S | 4506 hCoV-19/England/QEUH-AAC1DC/2020\|EPI_ISL_652847\|20 | ···················· | ···················· | ···················· | ···················· | ···················· | ···················· |
| S | 4507 hCoV-19/England/QEUH-AAC1FA/2020\|EPI_ISL_652848\|20 | ···················· | ···················· | ···················· | ···················· | ···················· | ···················· |
| S | 4508 hCoV-19/England/QEUH-AAC206/2020\|EPI_ISL_652849\|20 | ···················· | ···················· | ···················· | ···················· | ···················· | ···················· |
| S | 4509 hCoV-19/England/QEUH-AAC224/2020\|EPI_ISL_652850\|20 | ···················· | ···················· | ···················· | ···················· | ···················· | ···················· |
| S | 4510 hCoV-19/England/QEUH-AAC28E/2020\|EPI_ISL_652851\|20 | ···················· | ···················· | ···················· | ···················· | ···················· | ···················· |
| S | 4511 hCoV-19/England/QEUH-AAC813/2020\|EPI_ISL_652874\|20 | ···················· | ···················· | ···················· | ···················· | ···················· | ···················· |
| S | 4512 hCoV-19/England/QEUH-AAC831/2020\|EPI_ISL_652875\|20 | ···················· | ···················· | ···················· | ···················· | ···················· | ···················· |
| S | 4513 hCoV-19/Scotland/QEUH-AAC840/2020\|EPI_ISL_652876\|2 | ···················· | ···················· | ···················· | ···················· | ···················· | ···················· |
| S | 4514 hCoV-19/England/QEUH-AAC85F/2020\|EPI_ISL_652877\|20 | ···················· | ···················· | ···················· | ···················· | ···················· | ···················· |
| S | 4515 hCoV-19/England/QEUH-AAC86E/2020\|EPI_ISL_652878\|20 | ···················· | ·A·················· | ···················· | ···················· | ···················· | ···················· |
| S | 4516 hCoV-19/England/QEUH-AAC88C/2020\|EPI_ISL_652879\|20 | ···················· | ···················· | ···················· | ···················· | ···················· | ···················· |
| S | 4517 hCoV-19/England/QEUH-AAC8C8/2020\|EPI_ISL_652880\|20 | ···················· | ···················· | ···················· | ···················· | ···T················ | ···················· |
| S | 4518 hCoV-19/England/QEUH-AAC8F5/2020\|EPI_ISL_652881\|20 | ···················· | ···················· | ···················· | ···················· | ···················· | ···················· |

| | | | | | | | |
|---|---|---|---|---|---|---|---|
| S | 4519 hCoV-19/England/QEUH-AAC910/2020\|EPI_ISL_652882\|20 | ···················· | ···················· | ····················· | ···················· | ···················· | ···················· |
| S | 4520 hCoV-19/England/QEUH-AAC92F/2020\|EPI_ISL_652883\|20 | ···················· | ···················· | ····················· | ···················· | ···················· | ···················· |
| S | 4521 hCoV-19/Scotland/QEUH-AAC9F2/2020\|EPI_ISL_652884\|2 | ···················· | ···················· | ····················· | ···················· | ···················· | ···················· |
| S | 4522 hCoV-19/Scotland/QEUH-AACAA4/2020\|EPI_ISL_652890\|2 | ···················· | ···················· | ····················· | ···················· | ···················· | ···················· |
| S | 4523 hCoV-19/Scotland/QEUH-AACB1A/2020\|EPI_ISL_652896\|2 | ···················· | ···················· | ····················· | ···················· | ···················· | ···················· |
| S | 4524 hCoV-19/Scotland/QEUH-AACB38/2020\|EPI_ISL_652897\|2 | ···················· | ···················· | ····················· | ···················· | ···················· | ···················· |
| S | 4525 hCoV-19/England/QEUH-AACB47/2020\|EPI_ISL_652898\|20 | ···················· | ···················· | ····················· | ···················· | ···················· | ···················· |
| S | 4526 hCoV-19/England/QEUH-AACB56/2020\|EPI_ISL_652899\|20 | ···················· | ···················· | ····················· | ···················· | ···················· | ···················· |
| S | 4527 hCoV-19/USA/LA-EVTL698/2020\|EPI_ISL_653390\|2020-07 | ···················· | ······A············· | ····················· | ···················· | ···················· | ···················· |
| S | 4528 hCoV-19/USA/LA-EVTL699/2020\|EPI_ISL_653391\|2020-07 | ···················· | ·············A······ | ····················· | ···················· | ···················· | ···················· |
| S | 4529 hCoV-19/USA/LA-EVTL700/2020\|EPI_ISL_653392\|2020-07 | ···················· | ···················· | ····················· | ···················· | ···················· | ···················· |
| S | 4530 hCoV-19/USA/LA-EVTL701/2020\|EPI_ISL_653393\|2020-07 | ···················· | ···················· | ····················· | ···················· | ···················· | ···················· |
| S | 4531 hCoV-19/USA/LA-EVTL702/2020\|EPI_ISL_653394\|2020-07 | ···················· | ···················· | ····················· | ···················· | ···················· | ···················· |
| S | 4532 hCoV-19/USA/LA-EVTL703/2020\|EPI_ISL_653395\|2020-07 | ···················· | ···················· | ····················· | ···················· | ···················· | ···················· |
| S | 4533 hCoV-19/USA/LA-EVTL704/2020\|EPI_ISL_653396\|2020-07 | ···················· | ···················· | ····················· | ···················· | ···················· | ···················· |
| S | 4534 hCoV-19/USA/LA-EVTL705/2020\|EPI_ISL_653397\|2020-07 | ···················· | ···················· | ····················· | ···················· | ···················· | ···················· |
| S | 4535 hCoV-19/USA/LA-EVTL706/2020\|EPI_ISL_653398\|2020-07 | ···················· | ···················· | ····················· | ···················· | ···················· | ···················· |
| S | 4536 hCoV-19/USA/LA-EVTL707/2020\|EPI_ISL_653399\|2020-07 | ···················· | ···················· | ····················· | ···················· | ···················· | ···················· |
| S | 4537 hCoV-19/England/QEUH-B66A0F/2020\|EPI_ISL_655051\|20 | ···················· | ···················· | ····················· | ···················· | ······T············· | ···················· |
| S | 4538 hCoV-19/England/QEUH-B68C52/2020\|EPI_ISL_655053\|20 | ···················· | ···················· | ····················· | ···················· | ···················· | ···················· |
| S | 4539 hCoV-19/England/QEUH-B6AC32/2020\|EPI_ISL_655054\|20 | ···················· | ···················· | ····················· | ···················· | ···················· | ···················· |
| S | 4540 hCoV-19/England/QEUH-B669F3/2020\|EPI_ISL_655055\|20 | ···················· | ···················· | ····················· | ···················· | ···················· | ···················· |
| S | 4541 hCoV-19/England/QEUH-B6C91B/2020\|EPI_ISL_655056\|20 | ···················· | ···················· | ····················· | ···················· | ···················· | ···················· |
| S | 4542 hCoV-19/England/QEUH-B69E79/2020\|EPI_ISL_655057\|20 | ···················· | ···················· | ····················· | ···················· | ···················· | ···················· |
| S | 4543 hCoV-19/England/QEUH-B682D8/2020\|EPI_ISL_655059\|20 | ···················· | ···················· | ····················· | ···················· | ···················· | ···················· |
| S | 4544 hCoV-19/England/QEUH-B6C08F/2020\|EPI_ISL_655060\|20 | ···················· | ···················· | ····················· | ···················· | ···················· | ···················· |
| S | 4545 hCoV-19/England/QEUH-B68B91/2020\|EPI_ISL_655061\|20 | ···················· | ···················· | ····················· | ···················· | ···················· | ···················· |
| S | 4546 hCoV-19/England/QEUH-B69E5B/2020\|EPI_ISL_655062\|20 | ···················· | ···················· | ····················· | ···················· | ···················· | ···················· |
| S | 4547 hCoV-19/England/QEUH-B684E1/2020\|EPI_ISL_655064\|20 | ···················· | ···················· | ····················· | ···················· | ···················· | ···················· |
| S | 4548 hCoV-19/England/QEUH-B69644/2020\|EPI_ISL_655065\|20 | ···················· | ···················· | ····················· | ···················· | ···················· | ···················· |
| S | 4549 hCoV-19/England/QEUH-B67D9C/2020\|EPI_ISL_655066\|20 | ···················· | ···················· | ····················· | ···················· | ···················· | ···················· |
| S | 4550 hCoV-19/England/QEUH-B69565/2020\|EPI_ISL_655067\|20 | ···················· | ···················· | ····················· | ···················· | ···················· | ···················· |
| S | 4551 hCoV-19/England/QEUH-B6AC05/2020\|EPI_ISL_655069\|20 | ···················· | ···················· | ····················· | ···················· | ···················· | ···················· |
| S | 4552 hCoV-19/England/QEUH-B68557/2020\|EPI_ISL_655070\|20 | ···················· | ···················· | ····················· | ···················· | ···················· | ···················· |
| S | 4553 hCoV-19/England/QEUH-B6C359/2020\|EPI_ISL_655072\|20 | ···················· | ···················· | ····················· | ···················· | ···················· | ···················· |
| S | 4554 hCoV-19/England/QEUH-B6C0BC/2020\|EPI_ISL_655073\|20 | ···················· | ···················· | ····················· | ···················· | ···················· | ···················· |
| S | 4555 hCoV-19/England/QEUH-B687F7/2020\|EPI_ISL_655075\|20 | ···················· | ···················· | ····················· | ···················· | ···················· | ···················· |
| S | 4556 hCoV-19/England/QEUH-B6C83C/2020\|EPI_ISL_655076\|20 | ···················· | ···················· | ····················· | ···················· | ···················· | ···················· |
| S | 4557 hCoV-19/England/QEUH-B6BDE3/2020\|EPI_ISL_655078\|20 | ···················· | ·A·················· | ····················· | ···················· | ···················· | ···················· |
| S | 4558 hCoV-19/England/QEUH-B67CBD/2020\|EPI_ISL_655079\|20 | ···················· | ···················· | ····················· | ···················· | ···················· | ···················· |
| S | 4559 hCoV-19/England/QEUH-B6BE4A/2020\|EPI_ISL_655080\|20 | ···················· | ···················· | ····················· | ···················· | ···················· | ···················· |
| S | 4560 hCoV-19/England/QEUH-B6C395/2020\|EPI_ISL_655081\|20 | ···················· | ···················· | ····················· | ···················· | ···················· | ···················· |
| S | 4561 hCoV-19/England/QEUH-B6C4CF/2020\|EPI_ISL_655082\|20 | ···················· | ···················· | ····················· | ···················· | ···················· | ···················· |
| S | 4562 hCoV-19/England/QEUH-B65DBC/2020\|EPI_ISL_655083\|20 | ···················· | ···················· | ····················· | ···················· | ···················· | ···················· |
| S | 4563 hCoV-19/England/QEUH-B6C90C/2020\|EPI_ISL_655084\|20 | ···················· | ···················· | ····················· | ···················· | ···················· | ···················· |
| S | 4564 hCoV-19/England/QEUH-B6A25E/2020\|EPI_ISL_655085\|20 | ···················· | ···················· | ····················· | ···················· | ···················· | ···················· |
| S | 4565 hCoV-19/England/QEUH-B68AFE/2020\|EPI_ISL_655087\|20 | ···················· | ···················· | ····················· | ···················· | ···················· | ···················· |
| S | 4566 hCoV-19/England/QEUH-B6889A/2020\|EPI_ISL_655088\|20 | ···················· | ···················· | ····················· | ···················· | ···················· | ···················· |
| S | 4567 hCoV-19/England/QEUH-B68C16/2020\|EPI_ISL_655090\|20 | ···················· | ···················· | ····················· | ···················· | ···················· | ···················· |
| S | 4568 hCoV-19/England/QEUH-B68803/2020\|EPI_ISL_655091\|20 | ···················· | ···················· | ····················· | ···················· | ···················· | ···················· |
| S | 4569 hCoV-19/England/QEUH-B69C06/2020\|EPI_ISL_655092\|20 | ···················· | ···················· | ····················· | ···················· | ···················· | ···················· |
| S | 4570 hCoV-19/England/QEUH-B6951A/2020\|EPI_ISL_655093\|20 | ···················· | ···················· | ····················· | ···················· | ···················· | ···················· |
| S | 4571 hCoV-19/England/QEUH-B66A3C/2020\|EPI_ISL_655094\|20 | ···················· | ···················· | ····················· | ···················· | ···················· | ···················· |
| S | 4572 hCoV-19/England/QEUH-B68CCB/2020\|EPI_ISL_655095\|20 | ···················· | ···················· | ····················· | ···················· | ···················· | ···················· |
| S | 4573 hCoV-19/England/QEUH-B6B9A3/2020\|EPI_ISL_655096\|20 | ···················· | ···················· | ····················· | ···················· | ···················· | ···················· |
| S | 4574 hCoV-19/England/QEUH-B687E8/2020\|EPI_ISL_655097\|20 | ···················· | ···················· | ····················· | ···················· | ···················· | ···················· |
| S | 4575 hCoV-19/England/QEUH-B6850C/2020\|EPI_ISL_655099\|20 | ···················· | ···················· | ····················· | ···················· | ···················· | ···················· |
| S | 4576 hCoV-19/England/ALDP-B62696/2020\|EPI_ISL_656380\|20 | ···················· | ···················· | ····················· | ···················· | ···················· | ···················· |
| S | 4577 hCoV-19/England/ALDP-B638DB/2020\|EPI_ISL_656382\|20 | ···················· | ···················· | ····················· | ···················· | ···················· | ···················· |
| S | 4578 hCoV-19/England/ALDP-B63A3F/2020\|EPI_ISL_656383\|20 | ···················· | ···················· | ····················· | ···················· | ···················· | ···················· |
| S | 4579 hCoV-19/England/ALDP-B62809/2020\|EPI_ISL_656384\|20 | ···················· | ···················· | ····················· | ···················· | ···················· | ···················· |
| S | 4580 hCoV-19/England/ALDP-B625F3/2020\|EPI_ISL_656385\|20 | ···················· | ···················· | ····················· | ···················· | ···················· | ···················· |
| S | 4581 hCoV-19/England/ALDP-B62766/2020\|EPI_ISL_656386\|20 | ···················· | ···················· | ····················· | ···················· | ···················· | ···················· |
| S | 4582 hCoV-19/England/ALDP-B63B69/2020\|EPI_ISL_656388\|20 | ···················· | ···················· | ····················· | ···················· | ···················· | ···················· |
| S | 4583 hCoV-19/England/ALDP-B634D7/2020\|EPI_ISL_656389\|20 | ···················· | ···················· | ····················· | ···················· | ···················· | ···················· |
| S | 4584 hCoV-19/England/ALDP-B626C3/2020\|EPI_ISL_656391\|20 | ···················· | ···················· | ····················· | ···················· | ···················· | ···················· |
| S | 4585 hCoV-19/England/ALDP-B63826/2020\|EPI_ISL_656392\|20 | ···················· | ···················· | ····················· | ···················· | ···················· | ···················· |
| S | 4586 hCoV-19/England/ALDP-B61FF6/2020\|EPI_ISL_656393\|20 | ···················· | ···················· | ····················· | ···················· | ······T············· | ···················· |
| S | 4587 hCoV-19/England/ALDP-B63844/2020\|EPI_ISL_656394\|20 | ···················· | ···················· | ····················· | ···················· | ···················· | ···················· |
| S | 4588 hCoV-19/England/ALDP-B622ED/2020\|EPI_ISL_656395\|20 | ···················· | ···················· | ····················· | ···················· | ···················· | ···················· |
| S | 4589 hCoV-19/England/ALDP-B63352/2020\|EPI_ISL_656396\|20 | ···················· | ···················· | ····················· | ···················· | ···················· | ···················· |
| S | 4590 hCoV-19/England/ALDP-B63B78/2020\|EPI_ISL_656397\|20 | ···················· | ···················· | ····················· | ···················· | ···················· | ···················· |
| S | 4591 hCoV-19/England/ALDP-B6360E/2020\|EPI_ISL_656399\|20 | ···················· | ···················· | ····················· | ···················· | ···················· | ···················· |
| S | 4592 hCoV-19/England/CAMC-B343A5/2020\|EPI_ISL_659501\|20 | ···················· | ···················· | ····················· | ···················· | ···················· | ···················· |
| S | 4593 hCoV-19/England/CAMC-B329B4/2020\|EPI_ISL_659502\|20 | ···················· | ···················· | ····················· | ···················· | ···················· | ···················· |
| S | 4594 hCoV-19/England/CAMC-B35632/2020\|EPI_ISL_659503\|20 | ···················· | ···················· | ····················· | ···················· | ···················· | ···················· |
| S | 4595 hCoV-19/England/CAMC-B34026/2020\|EPI_ISL_659504\|20 | ···················· | ···················· | ····················· | ···················· | ···················· | ···················· |
| S | 4596 hCoV-19/England/CAMC-B3440C/2020\|EPI_ISL_659505\|20 | ···················· | ···················· | ····················· | ···················· | ···················· | ···················· |
| S | 4597 hCoV-19/England/CAMC-B32D8B/2020\|EPI_ISL_659507\|20 | ···················· | ···················· | ····················· | ···················· | ···················· | ···················· |
| S | 4598 hCoV-19/England/CAMC-B355BD/2020\|EPI_ISL_659508\|20 | ···················· | ···················· | ····················· | ···················· | ···················· | ···················· |

| | | | | | | | | |
|---|---|---|---|---|---|---|---|---|
| S | 4599 hCoV-19/England/CAMC-B350F8/2020\|EPI_ISL_659509\|20 | .................... | .................... | .................... | .................... | .................... | .................... | |
| S | 4600 hCoV-19/England/CAMC-B3285D/2020\|EPI_ISL_659518\|20 | .................... | .................... | .................... | .................... | .................... | .................... | |
| S | 4601 hCoV-19/England/CAMC-B34369/2020\|EPI_ISL_659519\|20 | .................... | .................... | .................... | .................... | .................... | .................... | |
| S | 4602 hCoV-19/England/CAMC-B3295A/2020\|EPI_ISL_659520\|20 | .................... | .................... | .................... | .................... | .................... | .................... | |
| S | 4603 hCoV-19/England/CAMC-B32A39/2020\|EPI_ISL_659522\|20 | .................... | .................... | .................... | .................... | .................... | .................... | |
| S | 4604 hCoV-19/England/CAMC-B3573F/2020\|EPI_ISL_659523\|20 | .................... | .................... | .................... | .................... | .................... | .................... | |
| S | 4605 hCoV-19/England/CAMC-B35474/2020\|EPI_ISL_659524\|20 | .................... | .................... | .................... | .................... | .................... | .................... | |
| S | 4606 hCoV-19/England/CAMC-B3575D/2020\|EPI_ISL_659526\|20 | .................... | .A.................. | .................... | .................... | .................... | ..A................. | |
| S | 4607 hCoV-19/England/CAMC-B34448/2020\|EPI_ISL_659527\|20 | .................... | .................... | .................... | .................... | .................... | .................... | |
| S | 4608 hCoV-19/England/CAMC-B3492B/2020\|EPI_ISL_659528\|20 | .................... | .................... | .................... | .................... | .................... | .................... | |
| S | 4612 hCoV-19/France/PDL-SC600/2020\|EPI_ISL_649173\|2020- | .................... | .................... | .................... | .................... | .................... | .................... | |
| S | 4613 hCoV-19/France/PDL-SC606/2020\|EPI_ISL_649174\|2020- | .................... | .................... | .................... | .................... | .................... | .................... | |
| S | 4614 hCoV-19/France/PDL-SC610/2020\|EPI_ISL_649175\|2020- | .................... | .................... | .................... | .................... | .................... | .................... | |
| S | 4615 hCoV-19/France/PDL-SC619/2020\|EPI_ISL_649176\|2020- | .................... | .................... | .................... | .................... | .................... | .................... | |
| S | 4616 hCoV-19/France/PDL-SC621/2020\|EPI_ISL_649177\|2020- | .................... | .................... | .................... | .................... | .................... | .................... | |
| S | 4617 hCoV-19/France/PDL-SC627/2020\|EPI_ISL_649178\|2020- | .................... | .................... | .................... | .................... | .................... | .................... | |
| S | 4618 hCoV-19/France/PDL-SC628/2020\|EPI_ISL_649179\|2020- | .................... | ...................A | .................... | .................... | .................... | .................... | 1 |
| S | 4619 hCoV-19/France/PDL-SC631/2020\|EPI_ISL_649180\|2020- | .................... | .................... | .................... | .................... | .................... | .................... | |
| S | 4620 hCoV-19/France/PDL-SC633/2020\|EPI_ISL_649181\|2020- | .................... | .................... | .................... | .................... | .................... | .................... | |
| S | 4621 hCoV-19/France/PDL-SC639/2020\|EPI_ISL_649182\|2020- | .................... | .................... | .................... | .................... | .................... | .................... | |
| S | 4622 hCoV-19/France/PDL-SC645/2020\|EPI_ISL_649183\|2020- | .................... | .................... | .................... | .................... | .................... | .................... | |
| S | 4623 hCoV-19/France/PDL-SC648/2020\|EPI_ISL_649184\|2020- | .................... | .................... | .................... | .................... | .................... | .................... | |
| S | 4624 hCoV-19/France/PDL-SC651/2020\|EPI_ISL_649185\|2020- | .................... | .................... | .................... | .................... | .................... | .................... | |
| S | 4625 hCoV-19/France/PDL-SC655/2020\|EPI_ISL_649186\|2020- | .................... | .................... | .................... | .................... | .................... | .................... | |
| S | 4626 hCoV-19/France/PDL-SC657/2020\|EPI_ISL_649187\|2020- | .................... | .................... | .................... | .................... | .................... | .................... | |
| S | 4627 hCoV-19/France/PAC-SC458/2020\|EPI_ISL_649188\|2020- | .................... | .................... | .................... | .................... | .................... | .................... | |
| S | 4628 hCoV-19/Italy/APU-IZSPB-98APT/2020\|EPI_ISL_649189\| | .................... | .................... | .................... | .................... | .................... | .................... | |
| S | 4629 hCoV-19/Italy/APU-IZSPB-170APT/2020\|EPI_ISL_649190 | .................... | .................... | .................... | .................... | .................... | .................... | |
| S | 4630 hCoV-19/Italy/APU-IZSPB-172APT/2020\|EPI_ISL_649191 | .................... | .................... | .................... | .................... | .................... | .................... | |
| S | 4631 hCoV-19/England/MILK-B50507/2020\|EPI_ISL_649192\|20 | .................... | .................... | .................... | .................... | ......T............. | .................... | |
| S | 4632 hCoV-19/England/ALDP-B48F6E/2020\|EPI_ISL_649193\|20 | .................... | .................... | .................... | .................... | .................... | .................... | |
| S | 4633 hCoV-19/England/ALDP-B48371/2020\|EPI_ISL_649194\|20 | .................... | .................... | .................... | .................... | .................... | .................... | |
| S | 4634 hCoV-19/England/ALDP-B483CC/2020\|EPI_ISL_649195\|20 | .................... | .................... | .................... | .................... | .................... | .................... | |
| S | 4635 hCoV-19/England/ALDP-B49413/2020\|EPI_ISL_649196\|20 | .................... | .................... | .................... | .................... | .................... | .................... | |
| S | 4636 hCoV-19/England/ALDP-B4810E/2020\|EPI_ISL_649197\|20 | .................... | .................... | .................... | .................... | .................... | .................... | |
| S | 4637 hCoV-19/England/ALDP-B48344/2020\|EPI_ISL_649198\|20 | .................... | .A.................. | .................... | .................... | .................... | .................... | |
| S | 4638 hCoV-19/England/ALDP-B47D0B/2020\|EPI_ISL_649199\|20 | .................... | .................... | .................... | .................... | .................... | .................... | |
| S | 4639 hCoV-19/England/ALDP-B48432/2020\|EPI_ISL_649210\|20 | .................... | .................... | .................... | .................... | .................... | .................... | |
| S | 4640 hCoV-19/England/ALDP-B4821A/2020\|EPI_ISL_649211\|20 | .................... | .................... | .................... | .................... | .................... | .................... | |
| S | 4641 hCoV-19/England/ALDP-B48E07/2020\|EPI_ISL_649212\|20 | .................... | .................... | .................... | .................... | .................... | .................... | |
| S | 4642 hCoV-19/England/ALDP-B49352/2020\|EPI_ISL_649213\|20 | .................... | .................... | .................... | .................... | .................... | .................... | |
| S | 4643 hCoV-19/England/ALDP-B49316/2020\|EPI_ISL_649214\|20 | .................... | .................... | .................... | .................... | .................... | .................... | |
| S | 4644 hCoV-19/England/ALDP-B493E9/2020\|EPI_ISL_649215\|20 | .................... | .................... | .................... | .................... | .................... | .................... | |
| S | 4645 hCoV-19/England/ALDP-B487B1/2020\|EPI_ISL_649216\|20 | .................... | .................... | .................... | .................... | .................... | .................... | |
| S | 4646 hCoV-19/England/ALDP-B4804D/2020\|EPI_ISL_649217\|20 | .................... | .................... | .................... | .................... | .................... | .................... | |
| S | 4647 hCoV-19/England/ALDP-B49422/2020\|EPI_ISL_649218\|20 | .................... | .A.................. | .................... | .................... | .................... | ..A................. | |
| S | 4648 hCoV-19/England/ALDP-B50419/2020\|EPI_ISL_649219\|20 | .................... | .................... | .................... | .................... | .................... | .................... | |
| S | 4649 hCoV-19/England/ALDP-B50ABD/2020\|EPI_ISL_649220\|20 | .................... | .................... | .................... | .................... | ......T............. | .................... | |
| S | 4650 hCoV-19/England/ALDP-B48353/2020\|EPI_ISL_649221\|20 | .................... | .................... | .................... | .................... | .................... | .................... | |
| S | 4651 hCoV-19/England/ALDP-B486B4/2020\|EPI_ISL_649222\|20 | .................... | .................... | .................... | .................... | .................... | .................... | |
| S | 4652 hCoV-19/England/ALDP-B48FAA/2020\|EPI_ISL_649223\|20 | .................... | .................... | .................... | .................... | .................... | .................... | |
| S | 4653 hCoV-19/England/ALDP-B483BD/2020\|EPI_ISL_649224\|20 | .................... | .................... | .................... | .................... | .................... | .................... | |
| S | 4654 hCoV-19/England/ALDP-B4903D/2020\|EPI_ISL_649226\|20 | .................... | .................... | .................... | .................... | .................... | .................... | |
| S | 4655 hCoV-19/England/ALDP-B49404/2020\|EPI_ISL_649227\|20 | .................... | .................... | .................... | .................... | .................... | .................... | |
| S | 4656 hCoV-19/England/ALDP-B503C1/2020\|EPI_ISL_649228\|20 | .................... | .................... | .................... | .................... | .................... | .................... | |
| S | 4657 hCoV-19/England/ALDP-B47D92/2020\|EPI_ISL_649229\|20 | .................... | .................... | .................... | .................... | .................... | .................... | |
| S | 4658 hCoV-19/England/ALDP-B483EA/2020\|EPI_ISL_649240\|20 | .................... | .................... | .................... | .................... | .................... | .................... | |
| S | 4659 hCoV-19/England/ALDP-B48784/2020\|EPI_ISL_649241\|20 | .................... | .................... | .................... | .................... | .................... | .................... | |
| S | 4660 hCoV-19/England/ALDP-B47DED/2020\|EPI_ISL_649242\|20 | .................... | .................... | .................... | .................... | .................... | .................... | |
| S | 4661 hCoV-19/England/ALDP-B4945F/2020\|EPI_ISL_649243\|20 | .................... | .................... | .................... | .................... | .................... | .................... | |
| S | 4662 hCoV-19/England/ALDP-B4847E/2020\|EPI_ISL_649246\|20 | .................... | .................... | .................... | .................... | .................... | .................... | |
| S | 4663 hCoV-19/England/ALDP-B48FB9/2020\|EPI_ISL_649247\|20 | .................... | .................... | .................... | .................... | .................... | .................... | |
| S | 4664 hCoV-19/England/ALDP-B47F50/2020\|EPI_ISL_649248\|20 | .................... | .................... | .................... | .................... | .................... | .................... | |
| S | 4665 hCoV-19/England/ALDP-B49000/2020\|EPI_ISL_649249\|20 | .................... | .................... | .................... | .................... | .................... | .................... | |
| S | 4666 hCoV-19/Scotland/EDB8660/2020\|EPI_ISL_651521\|2020- | .................... | .................... | .................... | .................... | .................... | .................... | |
| S | 4667 hCoV-19/Scotland/EDB8661/2020\|EPI_ISL_651522\|2020- | .................... | .................... | .................... | .................... | .................... | .................... | |
| S | 4670 hCoV-19/England/MILK-ACFD75/2020\|EPI_ISL_651525\|20 | .................... | .................... | .................... | .................... | .................... | .................... | |
| S | 4671 hCoV-19/England/MILK-B06BA4/2020\|EPI_ISL_651526\|20 | .................... | .A.................. | .................... | .................... | .................... | ..A................. | |
| S | 4672 hCoV-19/England/MILK-B39ABA/2020\|EPI_ISL_651527\|20 | .................... | .................... | .................... | .................... | .................... | .................... | |
| S | 4673 hCoV-19/Scotland/QEUH-A9CF6C/2020\|EPI_ISL_651528\|2 | .................... | .................... | .................... | .................... | .................... | .................... | |
| S | 4674 hCoV-19/Scotland/QEUH-AAB1CE/2020\|EPI_ISL_651529\|2 | .................... | .................... | .................... | .................... | .................... | .................... | |
| S | 4675 hCoV-19/England/SHEF-C4B25/2020\|EPI_ISL_651530\|202 | .................... | .A.................. | .................... | .................... | .................... | ..A................. | |
| S | 4676 hCoV-19/England/CAMB-1B88D1/2020\|EPI_ISL_651542\|20 | .................... | .................... | .................... | .................... | .................... | .................... | |
| S | 4677 hCoV-19/Scotland/QEUH-AAC400/2020\|EPI_ISL_652852\|2 | .................... | .................... | .................... | .................... | .................... | .................... | |
| S | 4678 hCoV-19/Scotland/QEUH-AAC42E/2020\|EPI_ISL_652853\|2 | .................... | .................... | .................... | .................... | .................... | .................... | |
| S | 4679 hCoV-19/England/QEUH-AAC43D/2020\|EPI_ISL_652854\|20 | .................... | .................... | .................... | .................... | .................... | .................... | |
| S | 4680 hCoV-19/Scotland/QEUH-AAC46A/2020\|EPI_ISL_652855\|2 | .................... | .................... | .................... | .................... | .................... | .................... | |
| S | 4681 hCoV-19/England/QEUH-AAC488/2020\|EPI_ISL_652856\|20 | .................... | .................... | .................... | .................... | .................... | .................... | |
| S | 4682 hCoV-19/England/QEUH-AAC5C1/2020\|EPI_ISL_652857\|20 | .................... | .................... | .................... | .................... | .................... | .................... | |
| S | 4683 hCoV-19/England/QEUH-AAC5FE/2020\|EPI_ISL_652858\|20 | .................... | .................... | .................... | .................... | .................... | .................... | |

| | | | | | | | |
|---|---|---|---|---|---|---|---|
| S | 4684 hCoV-19/England/QEUH-AAC60A/2020\|EPI_ISL_652859\|20 | ···················· | ···················· | ···················· | ···················· | ···················· | ···················· |
| S | 4685 hCoV-19/Scotland/QEUH-AAC619/2020\|EPI_ISL_652860\|2 | ···················· | ···················· | ···················· | ···················· | ···················· | ···················· |
| S | 4686 hCoV-19/England/QEUH-AAC628/2020\|EPI_ISL_652861\|20 | ···················· | ···················· | ···················· | ···················· | ···················· | ···················· |
| S | 4687 hCoV-19/England/QEUH-AAC637/2020\|EPI_ISL_652862\|20 | ···················· | ···················· | ···················· | ···················· | ···················· | ···················· |
| S | 4688 hCoV-19/England/QEUH-AAC646/2020\|EPI_ISL_652863\|20 | ···················· | ···················· | ···················· | ···················· | ···················· | ···················· |
| S | 4689 hCoV-19/England/QEUH-AAC655/2020\|EPI_ISL_652864\|20 | ···················· | ···················· | ···················· | ···················· | ···················· | ···················· |
| S | 4690 hCoV-19/England/QEUH-AAC6BF/2020\|EPI_ISL_652865\|20 | ···················· | ···················· | ···················· | ···················· | ···················· | ···················· |
| S | 4691 hCoV-19/England/QEUH-AAC6DD/2020\|EPI_ISL_652866\|20 | ···················· | ···················· | ···················· | ···················· | ···················· | ···················· |
| S | 4692 hCoV-19/England/QEUH-AAC6EC/2020\|EPI_ISL_652867\|20 | ···················· | ···················· | ···················· | ···················· | ···················· | ···················· |
| S | 4693 hCoV-19/England/QEUH-AAC6FB/2020\|EPI_ISL_652868\|20 | ···················· | ···················· | ···················· | ···················· | ···················· | ···················· |
| S | 4694 hCoV-19/England/QEUH-AAC707/2020\|EPI_ISL_652869\|20 | ···················· | ···················· | ···················· | ···················· | ···················· | ···················· |
| S | 4695 hCoV-19/England/QEUH-AAC725/2020\|EPI_ISL_652870\|20 | ···················· | ···················· | ···················· | ···················· | ···················· | ···················· |
| S | 4696 hCoV-19/England/QEUH-AAC7AD/2020\|EPI_ISL_652871\|20 | ···················· | ···················· | ···················· | ···················· | ···················· | ···················· |
| S | 4697 hCoV-19/England/QEUH-AAC7BC/2020\|EPI_ISL_652872\|20 | ···················· | ···················· | ···················· | ···················· | ···················· | ···················· |
| S | 4698 hCoV-19/England/QEUH-AAC804/2020\|EPI_ISL_652873\|20 | ···················· | ···················· | ···················· | ···················· | ···················· | ···················· |
| S | 4699 hCoV-19/Scotland/QEUH-AACA0E/2020\|EPI_ISL_652885\|2 | ···················· | ···················· | ···················· | ···················· | ···················· | ···················· |
| S | 4700 hCoV-19/Scotland/QEUH-AACA3B/2020\|EPI_ISL_652886\|2 | ···················· | ···················· | ···················· | ···················· | ···················· | ···················· |
| S | 4701 hCoV-19/Scotland/QEUH-AACA4A/2020\|EPI_ISL_652887\|2 | ···················· | ···················· | ···················· | ···················· | ···················· | ···················· |
| S | 4702 hCoV-19/Scotland/QEUH-AACA68/2020\|EPI_ISL_652888\|2 | ···················· | ···················· | ···················· | ···················· | ···················· | ···················· |
| S | 4703 hCoV-19/Scotland/QEUH-AACA95/2020\|EPI_ISL_652889\|2 | ···················· | ···················· | ···················· | ···················· | ···················· | ···················· |
| S | 4704 hCoV-19/Scotland/QEUH-AACAB3/2020\|EPI_ISL_652891\|2 | ···················· | ···················· | ···················· | ···················· | ···················· | ···················· |
| S | 4705 hCoV-19/England/QEUH-AACAC2/2020\|EPI_ISL_652892\|20 | ···················· | ···················· | ···················· | ···················· | ···················· | ···················· |
| S | 4706 hCoV-19/Scotland/QEUH-AACAE0/2020\|EPI_ISL_652893\|2 | ···················· | ···················· | ···················· | ···················· | ···················· | ···················· |
| S | 4707 hCoV-19/Scotland/QEUH-AACAFF/2020\|EPI_ISL_652894\|2 | ···················· | ···················· | ···················· | ···················· | ···················· | ···················· |
| S | 4708 hCoV-19/Scotland/QEUH-AACB0B/2020\|EPI_ISL_652895\|2 | ···················· | ···················· | ···················· | ···················· | ···················· | ···················· |
| S | 4709 hCoV-19/England/QEUH-B6C92A/2020\|EPI_ISL_655008\|20 | ···················· | ···················· | ···················· | ···················· | ···················· | ···················· |
| S | 4710 hCoV-19/England/QEUH-B65D9E/2020\|EPI_ISL_655009\|20 | ···················· | ···················· | ···················· | ···················· | ···················· | ···················· |
| S | 4711 hCoV-19/England/QEUH-B69C33/2020\|EPI_ISL_655012\|20 | ···················· | ···················· | ···················· | ···················· | ···················· | ···················· |
| S | 4712 hCoV-19/England/QEUH-B667DB/2020\|EPI_ISL_655013\|20 | ···················· | ···················· | ···················· | ···················· | ···················· | ···················· |
| S | 4713 hCoV-19/England/QEUH-B68CAD/2020\|EPI_ISL_655014\|20 | ···················· | ···················· | ···················· | ···················· | ···················· | ···················· |
| S | 4714 hCoV-19/England/QEUH-B6B9B2/2020\|EPI_ISL_655015\|20 | ···················· | ···················· | ···················· | ···················· | ···················· | ···················· |
| S | 4715 hCoV-19/England/QEUH-B69FA3/2020\|EPI_ISL_655016\|20 | ···················· | ···················· | ···················· | ···················· | ···················· | ···················· |
| S | 4716 hCoV-19/England/QEUH-B695B0/2020\|EPI_ISL_655017\|20 | ···················· | ···················· | ···················· | ···················· | ···················· | ···················· |
| S | 4717 hCoV-19/England/QEUH-B66744/2020\|EPI_ISL_655018\|20 | ···················· | ···················· | ···················· | ···················· | ···················· | ···················· |
| S | 4718 hCoV-19/England/QEUH-B69EA6/2020\|EPI_ISL_655020\|20 | ···················· | ···················· | ···················· | ···················· | ···················· | ···················· |
| S | 4719 hCoV-19/England/QEUH-B68760/2020\|EPI_ISL_655021\|20 | ···················· | ···················· | ···················· | ···················· | ···················· | ···················· |
| S | 4720 hCoV-19/England/QEUH-B66735/2020\|EPI_ISL_655023\|20 | ···················· | ···················· | ···················· | ···················· | ···················· | ···················· |
| S | 4721 hCoV-19/England/QEUH-B6BE1D/2020\|EPI_ISL_655024\|20 | ···················· | ···················· | ···················· | ···················· | ···················· | ···················· |
| S | 4722 hCoV-19/England/QEUH-B66A96/2020\|EPI_ISL_655025\|20 | ···················· | ···················· | ···················· | ···················· | ···················· | ···················· |
| S | 4723 hCoV-19/England/QEUH-B69C60/2020\|EPI_ISL_655026\|20 | ···················· | ···················· | ···················· | ···················· | ···················· | ···················· |
| S | 4724 hCoV-19/England/QEUH-B68AEF/2020\|EPI_ISL_655027\|20 | ···················· | ···················· | ···················· | ···················· | ···················· | ···················· |
| S | 4725 hCoV-19/England/QEUH-B667AE/2020\|EPI_ISL_655028\|20 | ···················· | ···················· | ···················· | ···················· | ···················· | ···················· |
| S | 4726 hCoV-19/England/QEUH-B6BDA7/2020\|EPI_ISL_655029\|20 | ···················· | ···················· | ···················· | ···················· | ···················· | ···················· |
| S | 4727 hCoV-19/England/QEUH-B68548/2020\|EPI_ISL_655031\|20 | ···················· | ···················· | ···················· | ···················· | ···················· | ···················· |
| S | 4728 hCoV-19/England/QEUH-B6BA64/2020\|EPI_ISL_655032\|20 | ···················· | ···················· | ···················· | ···················· | ···················· | ···················· |
| S | 4729 hCoV-19/England/QEUH-B67DE7/2020\|EPI_ISL_655034\|20 | ···················· | ···················· | ···················· | ···················· | ···················· | ···················· |
| S | 4730 hCoV-19/England/QEUH-B69F1C/2020\|EPI_ISL_655035\|20 | ···················· | ···················· | ···················· | ···················· | ···················· | ···················· |
| S | 4731 hCoV-19/England/QEUH-B6BDC5/2020\|EPI_ISL_655039\|20 | ···················· | ···················· | ···················· | ···················· | ···················· | ···················· |
| S | 4732 hCoV-19/England/QEUH-B66999/2020\|EPI_ISL_655040\|20 | ···················· | ···················· | ···················· | ···················· | ···················· | ···················· |
| S | 4733 hCoV-19/England/QEUH-B667EA/2020\|EPI_ISL_655041\|20 | ···················· | ···················· | ···················· | ···················· | ···················· | ···················· |
| S | 4734 hCoV-19/England/QEUH-B684C3/2020\|EPI_ISL_655042\|20 | ···················· | ···················· | ···················· | ···················· | ···················· | ···················· |
| S | 4735 hCoV-19/England/QEUH-B66AE1/2020\|EPI_ISL_655043\|20 | ···················· | ···················· | ···················· | ···················· | ···················· | ···················· |
| S | 4736 hCoV-19/England/QEUH-B6851B/2020\|EPI_ISL_655045\|20 | ···················· | ···················· | ···················· | ···················· | ···················· | ···················· |
| S | 4737 hCoV-19/England/QEUH-B69556/2020\|EPI_ISL_655047\|20 | ···················· | ···················· | ···················· | ···················· | ···················· | ···················· |
| S | 4738 hCoV-19/England/QEUH-B6BEA4/2020\|EPI_ISL_655049\|20 | ···················· | ···················· | ···················· | ···················· | ···················· | ···················· |
| S | 4739 hCoV-19/England/ALDP-B45F25/2020\|EPI_ISL_656339\|20 | ···················· | ···················· | ···················· | ···················· | ···················· | ···················· |
| S | 4740 hCoV-19/England/ALDP-B46FBB/2020\|EPI_ISL_656340\|20 | ···················· | ···················· | ···················· | ···················· | ···················· | ···················· |
| S | 4741 hCoV-19/England/ALDP-B46C5A/2020\|EPI_ISL_656341\|20 | ···················· | ···················· | ···················· | ···················· | ···················· | ···················· |
| S | 4742 hCoV-19/England/ALDP-B46AE7/2020\|EPI_ISL_656343\|20 | ···················· | ···················· | ···················· | ···················· | ···················· | ···················· |
| S | 4743 hCoV-19/England/ALDP-B460E5/2020\|EPI_ISL_656344\|20 | ···················· | ···················· | ···················· | ···················· | ···················· | ···················· |
| S | 4744 hCoV-19/England/ALDP-B62362/2020\|EPI_ISL_656345\|20 | ············T······· | ···················· | ···················· | ···················· | ···················· | ···················· |
| S | 4745 hCoV-19/England/ALDP-B637ED/2020\|EPI_ISL_656346\|20 | ···················· | ···················· | ···················· | ···················· | ···················· | ···················· |
| S | 4746 hCoV-19/England/ALDP-B61FD8/2020\|EPI_ISL_656347\|20 | ···················· | ···················· | ···················· | ···················· | ···················· | ···················· |
| S | 4747 hCoV-19/England/ALDP-B628AF/2020\|EPI_ISL_656348\|20 | ···················· | ···················· | ···················· | ···················· | ···················· | ···················· |
| S | 4748 hCoV-19/England/ALDP-B6361D/2020\|EPI_ISL_656350\|20 | ···················· | ···················· | ···················· | ···················· | ···················· | ···················· |
| S | 4749 hCoV-19/England/ALDP-B623AE/2020\|EPI_ISL_656351\|20 | ···················· | ···················· | ···················· | ···················· | ···················· | ···················· |
| S | 4750 hCoV-19/England/ALDP-B62951/2020\|EPI_ISL_656352\|20 | ···················· | ···················· | ···················· | ···················· | ···················· | ···················· |
| S | 4751 hCoV-19/England/ALDP-B62001/2020\|EPI_ISL_656354\|20 | ···················· | ···················· | ···················· | ···················· | ······T············· | ···················· |
| S | 4752 hCoV-19/England/ALDP-B62A5E/2020\|EPI_ISL_656355\|20 | ···················· | ···················· | ···················· | ···················· | ···················· | ···················· |
| S | 4753 hCoV-19/England/ALDP-B63316/2020\|EPI_ISL_656356\|20 | ···················· | ···················· | ···················· | ···················· | ···················· | ···················· |
| S | 4754 hCoV-19/England/ALDP-B63219/2020\|EPI_ISL_656357\|20 | ···················· | ···················· | ···················· | ···················· | ···················· | ···················· |
| S | 4755 hCoV-19/England/ALDP-B631FE/2020\|EPI_ISL_656358\|20 | ···················· | ···················· | ···················· | ···················· | ···················· | ···················· |
| S | 4756 hCoV-19/England/ALDP-B6349B/2020\|EPI_ISL_656359\|20 | ···················· | ···················· | ···················· | ···················· | ···················· | ···················· |
| S | 4757 hCoV-19/England/ALDP-B63501/2020\|EPI_ISL_656362\|20 | ···················· | ···················· | ···················· | ···················· | ···················· | ···················· |
| S | 4758 hCoV-19/England/ALDP-B62AF4/2020\|EPI_ISL_656363\|20 | ···················· | ···················· | ···················· | ···················· | ···················· | ···················· |
| S | 4759 hCoV-19/England/ALDP-B6298E/2020\|EPI_ISL_656364\|20 | ···················· | ···················· | ···················· | ···················· | ···················· | ···················· |
| S | 4760 hCoV-19/England/ALDP-B63B96/2020\|EPI_ISL_656365\|20 | ···················· | ···················· | ···················· | ···················· | ···················· | ···················· |
| S | 4761 hCoV-19/England/ALDP-B634E6/2020\|EPI_ISL_656366\|20 | ···················· | ···················· | ···················· | ···················· | ···················· | ···················· |
| S | 4762 hCoV-19/England/ALDP-B61F50/2020\|EPI_ISL_656367\|20 | ···················· | ···················· | ···················· | ···················· | ···················· | ···················· |
| S | 4763 hCoV-19/England/ALDP-B62A6D/2020\|EPI_ISL_656368\|20 | ···················· | ···················· | ···················· | ···················· | ···················· | ···················· |

S 4764 hCoV-19/England/ALDP-B629CA/2020|EPI_ISL_656369|20 ···················· ···················· ···················· ···················· ···················· ····················
S 4765 hCoV-19/England/ALDP-B63A02/2020|EPI_ISL_656370|20 ···················· ···················· ···················· ···················· ···················· ····················
S 4766 hCoV-19/England/ALDP-B62A12/2020|EPI_ISL_656371|20 ···················· ···················· ···················· ···················· ···················· ····················
S 4767 hCoV-19/England/ALDP-B63B1E/2020|EPI_ISL_656372|20 ···················· ···················· ···················· ···················· ···················· ····················
S 4768 hCoV-19/England/ALDP-B62827/2020|EPI_ISL_656374|20 ···················· ···················· ···················· ···················· ···················· ····················
S 4769 hCoV-19/England/ALDP-B63598/2020|EPI_ISL_656375|20 ···················· ···················· ···················· ···················· ···················· ····················
S 4770 hCoV-19/England/ALDP-B623EA/2020|EPI_ISL_656376|20 ···················· ···················· ···················· ···················· ···················· ····················
S 4771 hCoV-19/England/ALDP-B6249C/2020|EPI_ISL_656377|20 ···················· ···················· ···················· ···················· ···················· ····················
S 4772 hCoV-19/England/ALDP-B62441/2020|EPI_ISL_656378|20 ···················· ···················· ···················· ···················· ···················· ····················
S 4773 hCoV-19/England/ALDP-B63A99/2020|EPI_ISL_656379|20 ···················· ···················· ···················· ···················· ···················· ····················
S 4774 hCoV-19/England/CAMC-B52A8E/2020|EPI_ISL_657600|20 ···················· ···················· ···················· ···················· ······T············· ···············A····
S 4775 hCoV-19/England/MILK-B565E3/2020|EPI_ISL_657602|20 ···················· ···················· ···················· ···················· ···················· ····················
S 4776 hCoV-19/England/CAMC-B52FF8/2020|EPI_ISL_657603|20 ···················· ···················· ···················· ···················· ······T············· ····················
S 4777 hCoV-19/England/CAMC-B52857/2020|EPI_ISL_657604|20 ···················· ···················· ···················· ···················· ······T············· ····················
S 4778 hCoV-19/England/MILK-B56237/2020|EPI_ISL_657605|20 ···················· ···················· ···················· ···················· ···················· ····················
S 4779 hCoV-19/England/MILK-B565F2/2020|EPI_ISL_657606|20 ···················· ···················· ···················· ···················· ···················· ····················
S 4780 hCoV-19/England/MILK-B55B00/2020|EPI_ISL_657608|20 ···················· ···················· ···················· ···················· ···················· ····················
S 4781 hCoV-19/England/MILK-B56668/2020|EPI_ISL_657609|20 ···················· ···················· ···················· ···················· ···················· ····················
S 4782 hCoV-19/England/MILK-B55766/2020|EPI_ISL_657610|20 ···················· ···················· ···················· ···················· ···················· ····················
S 4783 hCoV-19/England/MILK-B56510/2020|EPI_ISL_657611|20 ···················· ···················· ···················· ···················· ···················· ····················
S 4784 hCoV-19/England/CAMC-B52F52/2020|EPI_ISL_657612|20 ···················· ·A·················· ···················· ···················· ···················· ····················
S 4785 hCoV-19/England/CAMC-B52E0A/2020|EPI_ISL_657613|20 ···················· ···················· ···················· ···················· ···················· ····················
S 4786 hCoV-19/England/MILK-B562A0/2020|EPI_ISL_657614|20 ···················· ·A·················· ···················· ···················· ···················· ···A················
S 4787 hCoV-19/England/MILK-B556F0/2020|EPI_ISL_657615|20 ···················· ···················· ···················· ···················· ···················· ····················
S 4788 hCoV-19/England/CAMC-B52936/2020|EPI_ISL_657617|20 ···················· ···················· ···················· ···················· ······T············· ····················
S 4789 hCoV-19/England/CAMC-B52D49/2020|EPI_ISL_657618|20 ···················· ···················· ···················· ···················· ···················· ····················
S 4790 hCoV-19/England/CAMC-B52E82/2020|EPI_ISL_657619|20 ···················· ···················· ···················· ···················· ···················· ····················
S 4791 hCoV-19/England/CAMC-B52EBF/2020|EPI_ISL_657620|20 ···················· ···················· ···················· ···················· ······T············· ····················
S 4792 hCoV-19/England/CAMC-B531C4/2020|EPI_ISL_657621|20 ···················· ···················· ···················· ···················· ···················· ····················
S 4793 hCoV-19/England/CAMC-B529CD/2020|EPI_ISL_657623|20 ···················· ···················· ···················· ···················· ······T············· ····················
S 4794 hCoV-19/England/MILK-B5660E/2020|EPI_ISL_657625|20 ···················· ···················· ···················· ···················· ···················· ····················
S 4795 hCoV-19/England/CAMC-B53100/2020|EPI_ISL_657670|20 ···················· ···················· ···················· ···················· ···················· ····················
S 4796 hCoV-19/England/CAMC-B52875/2020|EPI_ISL_657671|20 ···················· ···················· ···················· ···················· ···················· ····················
S 4797 hCoV-19/England/CAMC-B528FD/2020|EPI_ISL_657672|20 ···················· ···················· ···················· ···················· ···················· ····················
S 4798 hCoV-19/England/CAMC-B5312E/2020|EPI_ISL_657673|20 ···················· ···················· ···················· ···················· ···················· ····················
S 4799 hCoV-19/England/CAMC-B5311F/2020|EPI_ISL_657674|20 ···················· ···················· ···················· ···················· ···················· ····················
S 4800 hCoV-19/England/CAMC-B530C7/2020|EPI_ISL_657675|20 ···················· ···················· ···················· ···················· ···················· ····················
S 4801 hCoV-19/England/MILK-B566D1/2020|EPI_ISL_657676|20 ···················· ·A·················· ···················· ···················· ···················· ···A················
S 4802 hCoV-19/England/CAMC-B52E28/2020|EPI_ISL_657677|20 ···················· ···················· ···················· ···················· ···················· ····················
S 4803 hCoV-19/England/CAMC-B5308B/2020|EPI_ISL_657679|20 ···················· ···················· ···················· ···················· ···················· ····················
S 4804 hCoV-19/England/MILK-B561EF/2020|EPI_ISL_657680|20 ···················· ···················· ···················· ···················· ···················· ····················
S 4805 hCoV-19/England/MILK-B566FF/2020|EPI_ISL_657681|20 ···················· ···················· ···················· ···················· ···················· ····················
S 4806 hCoV-19/England/MILK-B55933/2020|EPI_ISL_657682|20 ···················· ···················· ···················· ···················· ···················· ····················
S 4807 hCoV-19/England/CAMC-B52D1C/2020|EPI_ISL_657684|20 ···················· ···················· ···················· ···················· ···················· ····················
S 4808 hCoV-19/England/CAMC-B53188/2020|EPI_ISL_657685|20 ···················· ···················· ···················· ···················· ···················· ····················
S 4809 hCoV-19/England/CAMC-B53355/2020|EPI_ISL_657686|20 ···················· ···················· ···················· ···················· ···················· ····················
S 4810 hCoV-19/England/CAMC-B527B4/2020|EPI_ISL_657687|20 ···················· ···················· ···················· ···················· ···················· ····················
S 4811 hCoV-19/England/MILK-B564C8/2020|EPI_ISL_657688|20 ···················· ···················· ···················· ···················· ···················· ····················
S 4812 hCoV-19/England/CAMC-B52E91/2020|EPI_ISL_657689|20 ···················· ···················· ···················· ···················· ···················· ····················
S 4813 hCoV-19/England/CAMC-B52A33/2020|EPI_ISL_657690|20 ···················· ···················· ···················· ···················· ···················· ····················
S 4814 hCoV-19/England/CAMC-B52F9E/2020|EPI_ISL_657691|20 ···················· ···················· ···················· ···················· ···················· ····················
S 4815 hCoV-19/England/MILK-B557DF/2020|EPI_ISL_657694|20 ···················· ···················· ···················· ···················· ···················· ····················
S 4816 hCoV-19/England/MILK-B55A6D/2020|EPI_ISL_657696|20 ···················· ···················· ···················· ···················· ···················· ····················
S 4817 hCoV-19/England/CAMC-B52796/2020|EPI_ISL_657697|20 ···················· ···················· ···················· ···················· ···················· ····················
S 4818 hCoV-19/England/CAMC-B52A15/2020|EPI_ISL_657698|20 ···················· ···················· ···················· ···················· ···················· ····················
S 4819 hCoV-19/England/CAMC-B52D76/2020|EPI_ISL_657699|20 ···················· ···················· ···················· ···················· ···················· ····················
S 4820 hCoV-19/England/CAMC-B37DB3/2020|EPI_ISL_658900|20 ···················· ···················· ···················· ···················· ···················· ····················
S 4822 hCoV-19/England/MILK-B38B30/2020|EPI_ISL_658902|20 ···················· ···················· ···················· ···················· ···················· ····················
S 4823 hCoV-19/Mexico/QROO-InDRE-53/2020|EPI_ISL_658904|2 ···················· ···················· ···················· ···················· ···················· ····················
S 4824 hCoV-19/England/CAMC-B37DA4/2020|EPI_ISL_658905|20 ···················· ···················· ···················· ···················· ···················· ····················
S 4825 hCoV-19/England/MILK-B395D7/2020|EPI_ISL_658908|20 ···················· ···················· ···················· ···················· ···················· ····················
S 4826 hCoV-19/England/CAMC-B37F35/2020|EPI_ISL_658909|20 ···················· ···················· ···················· ···················· ······T············· ···············A····
S 4827 hCoV-19/England/MILK-B389CD/2020|EPI_ISL_658910|20 ···················· ···················· ···················· ···················· ···················· ····················
S 4828 hCoV-19/England/MILK-B39188/2020|EPI_ISL_658911|20 ···················· ···················· ···················· ···················· ···················· ····················
S 4829 hCoV-19/England/CAMC-B37C2F/2020|EPI_ISL_658912|20 ···················· ···················· ···················· ···················· ······T············· ····················
S 4830 hCoV-19/England/CAMC-B37C10/2020|EPI_ISL_658913|20 ···················· ···················· ···················· ···················· ···················· ····················
S 4831 hCoV-19/England/CAMC-B3820E/2020|EPI_ISL_658914|20 ···················· ···················· ···················· ···················· ···················· ····················
S 4832 hCoV-19/England/MILK-B38356/2020|EPI_ISL_658915|20 ···················· ···················· ···················· ···················· ···················· ····················
S 4833 hCoV-19/England/CAMC-B37E29/2020|EPI_ISL_658916|20 ···················· ···················· ···················· ···················· ···················· ····················
S 4834 hCoV-19/England/MILK-B386F3/2020|EPI_ISL_658918|20 ···················· ···················· ···················· ···················· ···················· ····················
S 4835 hCoV-19/England/MILK-B392FE/2020|EPI_ISL_658920|20 ···················· ···················· ···················· ···················· ···················· ····················
S 4836 hCoV-19/England/MILK-B3A59A/2020|EPI_ISL_658922|20 ···················· ···················· ···················· ···················· ···················· ····················
S 4837 hCoV-19/England/CAMC-B3817A/2020|EPI_ISL_658923|20 ···················· ···················· ···················· ···················· ···················· ····················
S 4838 hCoV-19/England/MILK-B3871E/2020|EPI_ISL_658924|20 ···················· ···················· ···················· ···················· ···················· ····················
S 4839 hCoV-19/England/CAMC-B37F53/2020|EPI_ISL_658925|20 ···················· ·A·················· ···················· ···················· ···················· ···A················
S 4840 hCoV-19/England/MILK-B38C4C/2020|EPI_ISL_658926|20 ···················· ···················· ···················· ···················· ···················· ····················
S 4841 hCoV-19/England/MILK-B3881B/2020|EPI_ISL_658927|20 ···················· ···················· ···················· ···················· ···················· ····················
S 4842 hCoV-19/England/MILK-B39513/2020|EPI_ISL_658928|20 ···················· ···················· ···················· ···················· ···················· ····················
S 4843 hCoV-19/England/MILK-B388EE/2020|EPI_ISL_658929|20 ···················· ···················· ···················· ···················· ···················· ····················
S 4844 hCoV-19/England/CAMC-B382D1/2020|EPI_ISL_658930|20 ···················· ···················· ···················· ···················· ···················· ····················

| | | | | | | | | |
|---|---|---|---|---|---|---|---|---|
| S | 4845 hCoV-19/England/CAMC-B37D68/2020\|EPI_ISL_658931\|20 | ···················· | ···················· | ···················· | ···················· | ···················· | ···················· | |
| S | 4846 hCoV-19/England/MILK-B38EA0/2020\|EPI_ISL_658932\|20 | ···················· | ···················· | ···················· | ···················· | ···················· | ···················· | |
| S | 4847 hCoV-19/England/CAMC-B37FCC/2020\|EPI_ISL_658933\|20 | ···················· | ·A·······A·········· | ···················· | ···················· | ···················· | ···················· | |
| S | 4848 hCoV-19/England/MILK-B389EB/2020\|EPI_ISL_658934\|20 | ···················· | ···················· | ···················· | ···················· | ···················· | ···················· | |
| S | 4849 hCoV-19/England/MILK-B3955F/2020\|EPI_ISL_658935\|20 | ···················· | ···················· | ···················· | ···················· | ···················· | ···················· | |
| S | 4850 hCoV-19/England/MILK-B38392/2020\|EPI_ISL_658937\|20 | ···················· | ···················· | ···················· | ···················· | ···················· | ···················· | |
| S | 4851 hCoV-19/England/MILK-B38532/2020\|EPI_ISL_658939\|20 | ···················· | ·A·················· | ···················· | ···················· | ···················· | ···A················ | |
| S | 4852 hCoV-19/England/MILK-B38918/2020\|EPI_ISL_658941\|20 | ···················· | ···················· | ···················· | ···················· | ···················· | ···················· | |
| S | 4853 hCoV-19/England/MILK-B388B1/2020\|EPI_ISL_658942\|20 | ···················· | ···················· | ···················· | ···················· | ···················· | ···················· | |
| S | 4854 hCoV-19/England/MILK-B393FB/2020\|EPI_ISL_658944\|20 | ···················· | ···················· | ···················· | ···················· | ···················· | ···················· | |
| S | 4855 hCoV-19/England/MILK-B391F1/2020\|EPI_ISL_658946\|20 | ···················· | ···················· | ···················· | ···················· | ···················· | ···················· | |
| S | 4856 hCoV-19/England/MILK-B3A21B/2020\|EPI_ISL_658947\|20 | ···················· | ···················· | ···················· | ···················· | ···················· | ···················· | |
| S | 4857 hCoV-19/England/CAMC-B37E38/2020\|EPI_ISL_658948\|20 | ···················· | ···················· | ···················· | ···················· | ···················· | ···················· | |
| S | 4858 hCoV-19/England/MILK-B387B4/2020\|EPI_ISL_658950\|20 | ···················· | ···················· | ···················· | ···················· | ···················· | ···················· | |
| S | 4859 hCoV-19/England/MILK-B3A5D6/2020\|EPI_ISL_658951\|20 | ···················· | ···················· | ···················· | ···················· | ···················· | ···················· | |
| S | 4860 hCoV-19/England/MILK-B3849F/2020\|EPI_ISL_658952\|20 | ···················· | ···················· | ···················· | ···················· | ······T············· | ···················· | |
| S | 4861 hCoV-19/England/MILK-B3930A/2020\|EPI_ISL_658953\|20 | ···················· | ···················· | ···················· | ···················· | ···················· | ···················· | |
| S | 4862 hCoV-19/England/CAMC-B3824A/2020\|EPI_ISL_658954\|20 | ···················· | ···················· | ···················· | ···················· | ···················· | ···················· | |
| S | 4863 hCoV-19/England/MILK-B39531/2020\|EPI_ISL_658955\|20 | ···················· | ···················· | ···················· | ···················· | ···················· | ···················· | |
| S | 4864 hCoV-19/England/MILK-B38936/2020\|EPI_ISL_658956\|20 | ···················· | ···················· | ···················· | ···················· | ···················· | ···················· | |
| S | 4865 hCoV-19/Belgium/ITM_C5189_200701416/2020\|EPI_ISL_6 | ···················· | ···················· | ···················· | ···················· | ···················· | ···················· | |
| S | 4867 hCoV-19/Equatorial Guinea/16160/2020\|EPI_ISL_64915 | ···················· | ···················· | ···················· | ···················· | ·NN················· | ···················· | |
| S | 4869 hCoV-19/Equatorial Guinea/33881/2020\|EPI_ISL_64916 | ···················· | ···················· | ···················· | ···················· | NNN················· | ···················· | 1 |
| S | 4870 hCoV-19/England/MILK-ACFBD5/2020\|EPI_ISL_651431\|20 | ···················· | ···················· | ···················· | ···················· | ···················· | ···················· | |
| S | 4871 hCoV-19/England/MILK-ACFBE4/2020\|EPI_ISL_651432\|20 | ···················· | ···················· | ···················· | ···················· | ···················· | ···················· | |
| S | 4872 hCoV-19/Wales/PHWC-489B64/2020\|EPI_ISL_651444\|2020 | ···················· | ···················· | ···················· | ···················· | ···················· | ···················· | |
| S | 4873 hCoV-19/Wales/PHWC-489C07/2020\|EPI_ISL_651445\|2020 | ···················· | ···················· | ···················· | ···················· | ···················· | ···················· | |
| S | 4874 hCoV-19/Wales/PHWC-489C61/2020\|EPI_ISL_651446\|2020 | ···················· | ···················· | ···················· | ···················· | ···················· | ···················· | |
| S | 4875 hCoV-19/Wales/PHWC-489D13/2020\|EPI_ISL_651447\|2020 | ···················· | ···················· | ···················· | ···················· | ···················· | ···················· | |
| S | 4876 hCoV-19/Wales/PHWC-489D9B/2020\|EPI_ISL_651448\|2020 | ···················· | ···················· | ···················· | ···················· | ···················· | ···················· | |
| S | 4877 hCoV-19/Wales/PHWC-489DC8/2020\|EPI_ISL_651449\|2020 | ···················· | ···················· | ···················· | ···················· | ···················· | ···················· | |
| S | 4878 hCoV-19/Wales/PHWC-489E6B/2020\|EPI_ISL_651450\|2020 | ···················· | ···················· | ···················· | ···················· | ···················· | ···················· | |
| S | 4879 hCoV-19/Wales/PHWC-489EB6/2020\|EPI_ISL_651451\|2020 | ···················· | ···················· | ···················· | ···················· | ···················· | ···················· | |
| S | 4880 hCoV-19/Wales/PHWC-489EF2/2020\|EPI_ISL_651452\|2020 | ···················· | ···················· | ···················· | ···················· | ···················· | ···················· | |
| S | 4881 hCoV-19/Wales/PHWC-48A028/2020\|EPI_ISL_651453\|2020 | ···················· | ···················· | ···················· | ···················· | ···················· | ···················· | |
| S | 4882 hCoV-19/Wales/PHWC-48A073/2020\|EPI_ISL_651454\|2020 | ···················· | ···················· | ···················· | ···················· | ···················· | ···················· | |
| S | 4883 hCoV-19/Wales/PHWC-48A0CE/2020\|EPI_ISL_651455\|2020 | ···················· | ···················· | ···················· | ···················· | ···················· | ···················· | |
| S | 4884 hCoV-19/Wales/PHWC-48A0EC/2020\|EPI_ISL_651456\|2020 | ···················· | ···················· | ···················· | ···················· | ···················· | ···················· | |
| S | 4885 hCoV-19/Wales/PHWC-48A134/2020\|EPI_ISL_651457\|2020 | ···················· | ···················· | ···················· | ···················· | ···················· | ···················· | |
| S | 4886 hCoV-19/Wales/PHWC-48A27D/2020\|EPI_ISL_651458\|2020 | ···················· | ···················· | ···················· | ···················· | ···················· | ···················· | |
| S | 4887 hCoV-19/Wales/PHWC-48A2C8/2020\|EPI_ISL_651459\|2020 | ···················· | ···················· | ···················· | ···················· | ···················· | ···················· | |
| S | 4888 hCoV-19/Wales/PHWC-48A41D/2020\|EPI_ISL_651460\|2020 | ···················· | ···················· | ···················· | ···················· | ···················· | ···················· | |
| S | 4889 hCoV-19/Wales/PHWC-48A44A/2020\|EPI_ISL_651461\|2020 | ···················· | ···················· | ···················· | ···················· | ···················· | ···················· | |
| S | 4890 hCoV-19/Wales/PHWC-48A477/2020\|EPI_ISL_651462\|2020 | ···················· | ···················· | ···················· | ···················· | ···················· | ···················· | |
| S | 4891 hCoV-19/Wales/PHWC-48A76F/2020\|EPI_ISL_651463\|2020 | ···················· | ···················· | ···················· | ···················· | ···················· | ···················· | |
| S | 4892 hCoV-19/Wales/PHWC-48A996/2020\|EPI_ISL_651464\|2020 | ···················· | ···················· | ···················· | ···················· | ···················· | ···················· | |
| S | 4893 hCoV-19/Wales/PHWC-48A9A5/2020\|EPI_ISL_651465\|2020 | ···················· | ···················· | ···················· | ···················· | ···················· | ···················· | |
| S | 4894 hCoV-19/England/QEUH-A9D9F4/2020\|EPI_ISL_651466\|20 | ···················· | ···················· | ···················· | ···················· | ···················· | ···················· | |
| S | 4895 hCoV-19/England/QEUH-A9FD7E/2020\|EPI_ISL_651467\|20 | ···················· | ···················· | ···················· | ···················· | ···················· | ···················· | |
| S | 4896 hCoV-19/England/QEUH-AA0035/2020\|EPI_ISL_651468\|20 | ···················· | ···················· | ···················· | ···················· | ···················· | ···················· | |
| S | 4897 hCoV-19/England/QEUH-AAAC46/2020\|EPI_ISL_651469\|20 | ···················· | ·A·················· | ···················· | ···················· | ···················· | ···A················ | |
| S | 4898 hCoV-19/England/QEUH-AAACBF/2020\|EPI_ISL_651470\|20 | ···················· | ···················· | ···················· | ···················· | ···················· | ···················· | |
| S | 4899 hCoV-19/England/QEUH-AAAF2F/2020\|EPI_ISL_651471\|20 | ···················· | ···················· | ···················· | ···················· | ···················· | ···················· | |
| S | 4900 hCoV-19/England/QEUH-AAB3E6/2020\|EPI_ISL_651472\|20 | ···················· | ···················· | ···················· | ···················· | ···················· | ···················· | |
| S | 4901 hCoV-19/England/QEUH-AAB6ED/2020\|EPI_ISL_651473\|20 | ···················· | ···················· | ···················· | ···················· | ···················· | ···················· | |
| S | 4902 hCoV-19/England/QEUH-AABA96/2020\|EPI_ISL_651474\|20 | ···················· | ···················· | ···················· | ···················· | ···················· | ···················· | |
| S | 4903 hCoV-19/England/QEUH-AAC664/2020\|EPI_ISL_651475\|20 | ···················· | ···················· | ···················· | ···················· | ···················· | ···················· | |
| S | 4904 hCoV-19/England/QEUH-AAC673/2020\|EPI_ISL_651476\|20 | ···················· | ···················· | ···················· | ···················· | ···················· | ···················· | |
| S | 4905 hCoV-19/England/QEUH-AADB37/2020\|EPI_ISL_651477\|20 | ···················· | ···················· | ···················· | ···················· | ···················· | ···················· | |
| S | 4906 hCoV-19/England/QEUH-AADE4D/2020\|EPI_ISL_651478\|20 | ···················· | ···················· | ···················· | ···················· | ···················· | ···················· | |
| S | 4907 hCoV-19/England/QEUH-AAE9B4/2020\|EPI_ISL_651479\|20 | ···················· | ···················· | ···················· | ···················· | ···················· | ···················· | |
| S | 4908 hCoV-19/England/QEUH-AAEA93/2020\|EPI_ISL_651480\|20 | ···················· | ···················· | ···················· | ···················· | ···················· | ···················· | |
| S | 4909 hCoV-19/England/QEUH-AAECCA/2020\|EPI_ISL_651481\|20 | ···················· | ···················· | ···················· | ···················· | ···················· | ···················· | |
| S | 4910 hCoV-19/Scotland/QEUH-AAF1CA/2020\|EPI_ISL_651482\|2 | ···················· | ···················· | ···················· | ···················· | ···················· | ···················· | |
| S | 4911 hCoV-19/Scotland/QEUH-AAF3D3/2020\|EPI_ISL_651483\|2 | ···················· | ···················· | ···················· | ···················· | ···················· | ···················· | |
| S | 4912 hCoV-19/England/QEUH-AF60C7/2020\|EPI_ISL_651484\|20 | ···················· | ···················· | ···················· | ···················· | ···················· | ···················· | |
| S | 4913 hCoV-19/England/BIRM-67B60/2020\|EPI_ISL_651486\|202 | ···················· | ···················· | ···················· | ···················· | ···················· | ···················· | |
| S | 4914 hCoV-19/England/QEUH-AA0017/2020\|EPI_ISL_652700\|20 | ···················· | ·A·················· | ···················· | ···················· | ···················· | ···················· | |
| S | 4915 hCoV-19/England/QEUH-AA0026/2020\|EPI_ISL_652701\|20 | ···················· | ···················· | ···················· | ···················· | ···················· | ···················· | |
| S | 4916 hCoV-19/England/QEUH-AA0044/2020\|EPI_ISL_652702\|20 | ···················· | ···················· | ···················· | ···················· | ···················· | ···················· | |
| S | 4917 hCoV-19/England/QEUH-AA0053/2020\|EPI_ISL_652703\|20 | ···················· | ···················· | ···················· | ···················· | ···················· | ···················· | |
| S | 4918 hCoV-19/England/QEUH-AA0071/2020\|EPI_ISL_652704\|20 | ···················· | ···················· | ···················· | ···················· | ···················· | ···················· | |
| S | 4919 hCoV-19/England/QEUH-AA009F/2020\|EPI_ISL_652705\|20 | ···················· | ···················· | ···················· | ···················· | ···················· | ···················· | |
| S | 4920 hCoV-19/England/QEUH-AA00AE/2020\|EPI_ISL_652706\|20 | ···················· | ···················· | ···················· | ···················· | ···················· | ···················· | |
| S | 4921 hCoV-19/England/QEUH-AA00CC/2020\|EPI_ISL_652707\|20 | ···················· | ···················· | ···················· | ···················· | ···················· | ···················· | |
| S | 4922 hCoV-19/England/QEUH-AA0114/2020\|EPI_ISL_652708\|20 | ···················· | ···················· | ···················· | ···················· | ···················· | ···················· | |
| S | 4923 hCoV-19/England/QEUH-AA04EE/2020\|EPI_ISL_652709\|20 | ···················· | ···················· | ···················· | ···················· | ···················· | ···················· | |
| S | 4924 hCoV-19/England/QEUH-AA05AF/2020\|EPI_ISL_652710\|20 | ···················· | ···················· | ···················· | ···················· | ···················· | ···················· | |
| S | 4925 hCoV-19/England/QEUH-AAA806/2020\|EPI_ISL_652711\|20 | ···················· | ···················· | ···················· | ···················· | ···················· | ···················· | |
| S | 4926 hCoV-19/England/QEUH-AAA824/2020\|EPI_ISL_652712\|20 | ···················· | ···················· | ···················· | ···················· | ···················· | ···················· | |

| | | | | | | | | |
|---|---|---|---|---|---|---|---|---|
| S | 4927 | hCoV-19/England/QEUH-AAA860/2020\|EPI_ISL_652713\|20 | ···················· | ···················· | ····················· | ···················· | ···················· | ···················· |
| S | 4928 | hCoV-19/England/QEUH-AAA88E/2020\|EPI_ISL_652714\|20 | ···················· | ···················· | ····················· | ···················· | ···················· | ···················· |
| S | 4929 | hCoV-19/England/QEUH-AAA89D/2020\|EPI_ISL_652715\|20 | ···················· | ···················· | ····················· | ···················· | ···················· | ···················· |
| S | 4930 | hCoV-19/England/QEUH-AAA8AC/2020\|EPI_ISL_652716\|20 | ···················· | ···················· | ····················· | ···················· | ···················· | ···················· |
| S | 4931 | hCoV-19/England/QEUH-AAA8CA/2020\|EPI_ISL_652717\|20 | ···················· | ···················· | ····················· | ···················· | ···················· | ···················· |
| S | 4932 | hCoV-19/England/QEUH-AAA8E8/2020\|EPI_ISL_652718\|20 | ···················· | ·A·················· | ····················· | ···················· | ···················· | ··A················· |
| S | 4933 | hCoV-19/England/QEUH-AAA921/2020\|EPI_ISL_652719\|20 | ···················· | ···················· | ····················· | ···················· | ···················· | ···················· |
| S | 4934 | hCoV-19/Scotland/QEUH-AAA930/2020\|EPI_ISL_652720\|2 | ···················· | ···················· | ····················· | ···················· | ···················· | ···················· |
| S | 4935 | hCoV-19/England/QEUH-AAA95E/2020\|EPI_ISL_652721\|20 | ···················· | ·A·················· | ····················· | ···················· | ···················· | ··A················· |
| S | 4936 | hCoV-19/England/QEUH-AAA97C/2020\|EPI_ISL_652722\|20 | ···················· | ···················· | ····················· | ···················· | ···················· | ···················· |
| S | 4937 | hCoV-19/England/QEUH-AAA99A/2020\|EPI_ISL_652723\|20 | ···················· | ···················· | ····················· | ···················· | ···················· | ···················· |
| S | 4938 | hCoV-19/England/QEUH-AAA9A9/2020\|EPI_ISL_652724\|20 | ···················· | ···················· | ····················· | ···················· | ···················· | ···················· |
| S | 4939 | hCoV-19/England/QEUH-AAA9B8/2020\|EPI_ISL_652725\|20 | ···················· | ···················· | ····················· | ···················· | ···················· | ···················· |
| S | 4940 | hCoV-19/England/QEUH-AAA9C7/2020\|EPI_ISL_652726\|20 | ···················· | ···················· | ····················· | ···················· | ···················· | ···················· |
| S | 4941 | hCoV-19/England/QEUH-AAA9D6/2020\|EPI_ISL_652727\|20 | ···················· | ···················· | ····················· | ···················· | ···················· | ···················· |
| S | 4942 | hCoV-19/England/QEUH-AAA9E5/2020\|EPI_ISL_652728\|20 | ···················· | ···················· | ····················· | ···················· | ···················· | ···················· |
| S | 4943 | hCoV-19/Scotland/QEUH-AAAA4C/2020\|EPI_ISL_652729\|2 | ···················· | ···················· | ····················· | ···················· | ···················· | ···················· |
| S | 4944 | hCoV-19/Scotland/QEUH-AAAA5B/2020\|EPI_ISL_652730\|2 | ···················· | ···················· | ····················· | ···················· | ···················· | ···················· |
| S | 4945 | hCoV-19/England/QEUH-AAAF89/2020\|EPI_ISL_652760\|20 | ···················· | ···················· | ····················· | ···················· | ···················· | ···················· |
| S | 4946 | hCoV-19/England/QEUH-AAAFA7/2020\|EPI_ISL_652762\|20 | ···················· | ···················· | ····················· | ···················· | ···················· | ···················· |
| S | 4947 | hCoV-19/England/QEUH-AAAFB6/2020\|EPI_ISL_652763\|20 | ···················· | ···················· | ····················· | ···················· | ···················· | ···················· |
| S | 4948 | hCoV-19/Scotland/QEUH-AAB155/2020\|EPI_ISL_652775\|2 | ···················· | ···················· | ····················· | ···················· | ···················· | ···················· |
| S | 4949 | hCoV-19/Scotland/QEUH-AAB173/2020\|EPI_ISL_652776\|2 | ···················· | ···················· | ····················· | ···················· | ···················· | ···················· |
| S | 4950 | hCoV-19/England/QEUH-AAB1EC/2020\|EPI_ISL_652777\|20 | ···················· | ···················· | ····················· | ···················· | ···················· | ···················· |
| S | 4951 | hCoV-19/England/QEUH-AAB1FB/2020\|EPI_ISL_652778\|20 | ···················· | G··················· | ····················· | ···················· | ···················· | ···················· |
| S | 4952 | hCoV-19/England/QEUH-AAB252/2020\|EPI_ISL_652779\|20 | ···················· | ···················· | ····················· | ···················· | ···················· | ···················· |
| S | 4953 | hCoV-19/England/QEUH-AAB29E/2020\|EPI_ISL_652780\|20 | ···················· | ···················· | ····················· | ···················· | ···················· | ···················· |
| S | 4954 | hCoV-19/England/QEUH-AAB2AD/2020\|EPI_ISL_652781\|20 | ···················· | ···················· | ····················· | ···················· | ···················· | ···················· |
| S | 4955 | hCoV-19/England/QEUH-AAB2CB/2020\|EPI_ISL_652782\|20 | ···················· | ·A·················· | ····················· | ···················· | ···················· | ···················· |
| S | 4956 | hCoV-19/England/QEUH-AAB2E9/2020\|EPI_ISL_652783\|20 | ···················· | ···················· | ····················· | ···················· | ···················· | ···················· |
| S | 4957 | hCoV-19/England/QEUH-AAB313/2020\|EPI_ISL_652784\|20 | ···················· | ·A·················· | ····················· | ···················· | ···················· | ···················· |
| S | 4958 | hCoV-19/England/QEUH-AAB322/2020\|EPI_ISL_652785\|20 | ···················· | ···················· | ····················· | ···················· | ···················· | ···················· |
| S | 4959 | hCoV-19/England/QEUH-AAB340/2020\|EPI_ISL_652786\|20 | ···················· | ···················· | ····················· | ···················· | ···················· | ···················· |
| S | 4960 | hCoV-19/England/QEUH-AAB37D/2020\|EPI_ISL_652787\|20 | ···················· | ···················· | ····················· | ···················· | ···················· | ···················· |
| S | 4961 | hCoV-19/England/QEUH-AAB3C8/2020\|EPI_ISL_652788\|20 | ···················· | ···················· | ····················· | ···················· | ···················· | ···················· |
| S | 4962 | hCoV-19/England/QEUH-AAB3D7/2020\|EPI_ISL_652789\|20 | ···················· | ···················· | ····················· | ···················· | ···················· | ···················· |
| S | 4963 | hCoV-19/England/QEUH-AAB410/2020\|EPI_ISL_652790\|20 | ···················· | ···················· | ····················· | ···················· | ···················· | ···················· |
| S | 4964 | hCoV-19/England/QEUH-AAB45C/2020\|EPI_ISL_652791\|20 | ···················· | ···················· | ····················· | ···················· | ···················· | ···················· |
| S | 4965 | hCoV-19/Scotland/QEUH-AAB46B/2020\|EPI_ISL_652792\|2 | ···················· | ···················· | ····················· | ···················· | ···················· | ···················· |
| S | 4966 | hCoV-19/Scotland/QEUH-AAB4B6/2020\|EPI_ISL_652793\|2 | ···················· | ···················· | ····················· | ···················· | ···················· | ···················· |
| S | 4967 | hCoV-19/England/QEUH-AAB4C5/2020\|EPI_ISL_652794\|20 | ···················· | ···················· | ····················· | ···················· | ···················· | ···················· |
| S | 4968 | hCoV-19/England/QEUH-AAB4D4/2020\|EPI_ISL_652795\|20 | ···················· | ···················· | ····················· | ···················· | ···················· | ···················· |
| S | 4969 | hCoV-19/Scotland/QEUH-AAB4E3/2020\|EPI_ISL_652796\|2 | ···················· | ···················· | ····················· | ···················· | ···················· | ···················· |
| S | 4970 | hCoV-19/Scotland/QEUH-AAB559/2020\|EPI_ISL_652797\|2 | ···················· | ···················· | ····················· | ···················· | ···················· | ···················· |
| S | 4971 | hCoV-19/England/QEUH-AAB577/2020\|EPI_ISL_652798\|20 | ···················· | ···················· | ····················· | ···················· | ···················· | ···················· |
| S | 4972 | hCoV-19/Scotland/QEUH-AAB5A4/2020\|EPI_ISL_652799\|2 | ···················· | ···················· | ····················· | ···················· | ···················· | ···················· |
| S | 4973 | hCoV-19/England/QEUH-B6887C/2020\|EPI_ISL_655001\|20 | ···················· | ···················· | ····················· | ···················· | ···················· | ···················· |
| S | 4974 | hCoV-19/England/QEUH-B6C8E1/2020\|EPI_ISL_655002\|20 | ···················· | ···················· | ····················· | ···················· | ···················· | ···················· |
| S | 4975 | hCoV-19/England/QEUH-B67D7E/2020\|EPI_ISL_655003\|20 | ···················· | ·A·················· | ····················· | ···················· | ···················· | ··A················· |
| S | 4976 | hCoV-19/England/QEUH-B6BDF2/2020\|EPI_ISL_655004\|20 | ···················· | ···················· | ····················· | ···················· | ···················· | ···················· |
| S | 4977 | hCoV-19/England/QEUH-B6AC8D/2020\|EPI_ISL_655005\|20 | ···················· | ···················· | ····················· | ···················· | ···················· | ···················· |
| S | 4978 | hCoV-19/England/QEUH-B68232/2020\|EPI_ISL_655006\|20 | ···················· | ···················· | ····················· | ···················· | ···················· | ···················· |
| S | 4979 | hCoV-19/England/QEUH-B66AD2/2020\|EPI_ISL_655007\|20 | ···················· | ···················· | ····················· | ···················· | ···················· | ···················· |
| S | 4980 | hCoV-19/England/QEUH-B6BA28/2020\|EPI_ISL_655019\|20 | ···················· | ···················· | ····················· | ···················· | ···················· | ···················· |
| S | 4981 | hCoV-19/England/ALDP-B4703F/2020\|EPI_ISL_656260\|20 | ···················· | ···················· | ····················· | ···················· | ···················· | ···················· |
| S | 4982 | hCoV-19/England/ALDP-B46BF3/2020\|EPI_ISL_656261\|20 | ···················· | ···················· | ····················· | ···················· | ···················· | ···················· |
| S | 4983 | hCoV-19/England/ALDP-B46197/2020\|EPI_ISL_656262\|20 | ···················· | ···················· | ····················· | ···················· | ···················· | ···················· |
| S | 4984 | hCoV-19/England/ALDP-B47011/2020\|EPI_ISL_656263\|20 | ···················· | ···················· | ····················· | ···················· | ···················· | ···················· |
| S | 4985 | hCoV-19/England/ALDP-B45EEC/2020\|EPI_ISL_656264\|20 | ···················· | ···················· | ····················· | ···················· | ···················· | ···················· |
| S | 4986 | hCoV-19/England/ALDP-B45B9A/2020\|EPI_ISL_656265\|20 | ···················· | ···················· | ····················· | ···················· | ···················· | ···················· |
| S | 4987 | hCoV-19/England/ALDP-B45D94/2020\|EPI_ISL_656266\|20 | ···················· | ·········T·········· | ····················· | ···················· | ···················· | ···················· |
| S | 4988 | hCoV-19/England/ALDP-B45B30/2020\|EPI_ISL_656267\|20 | ···················· | ···················· | ····················· | ···················· | ···················· | ···················· |
| S | 4989 | hCoV-19/England/ALDP-B45DB2/2020\|EPI_ISL_656268\|20 | ···················· | ···················· | ····················· | ···················· | ···················· | ···················· |
| S | 4990 | hCoV-19/England/ALDP-B4656E/2020\|EPI_ISL_656269\|20 | ···················· | ···················· | ····················· | ···················· | ···················· | ···················· |
| S | 4991 | hCoV-19/England/ALDP-B45BD6/2020\|EPI_ISL_656270\|20 | ···················· | ···················· | ····················· | ···················· | ···················· | ···················· |
| S | 4992 | hCoV-19/England/ALDP-B46373/2020\|EPI_ISL_656271\|20 | ···················· | ···················· | ····················· | ···················· | ···················· | ···················· |
| S | 4993 | hCoV-19/England/ALDP-B4658C/2020\|EPI_ISL_656272\|20 | ···················· | ···················· | ····················· | ···················· | ···················· | ···················· |
| S | 4994 | hCoV-19/England/ALDP-B469F9/2020\|EPI_ISL_656273\|20 | ···················· | ···················· | ····················· | ···················· | ···················· | ···················· |
| S | 4995 | hCoV-19/England/ALDP-B45E0A/2020\|EPI_ISL_656274\|20 | ···················· | ···················· | ····················· | ···················· | ···················· | ···················· |
| S | 4996 | hCoV-19/England/ALDP-B46276/2020\|EPI_ISL_656276\|20 | ···················· | ···················· | ····················· | ···················· | ···················· | ···················· |
| S | 4997 | hCoV-19/England/ALDP-B46601/2020\|EPI_ISL_656277\|20 | ···················· | ···················· | ····················· | ···················· | ···················· | ···················· |
| S | 4998 | hCoV-19/England/ALDP-B465AA/2020\|EPI_ISL_656278\|20 | ···················· | ···················· | ····················· | ···················· | ···················· | ···················· |
| S | 4999 | hCoV-19/England/ALDP-B45AD9/2020\|EPI_ISL_656279\|20 | ···················· | ···················· | ····················· | ···················· | ···················· | ···················· |
| S | 5000 | hCoV-19/England/ALDP-B45EFB/2020\|EPI_ISL_656280\|20 | ···················· | ···················· | ····················· | ···················· | ···················· | ···················· |
| S | 5001 | hCoV-19/England/ALDP-B46D0C/2020\|EPI_ISL_656281\|20 | ···················· | ···················· | ····················· | ···················· | ···················· | ···················· |
| S | 5002 | hCoV-19/England/ALDP-B46267/2020\|EPI_ISL_656282\|20 | ···················· | ···················· | ····················· | ···················· | ···················· | ···················· |
| S | 5003 | hCoV-19/England/ALDP-B46B4E/2020\|EPI_ISL_656283\|20 | ···················· | ···················· | ····················· | ···················· | ···················· | ··A················· |
| S | 5004 | hCoV-19/England/ALDP-B45BC7/2020\|EPI_ISL_656284\|20 | ···················· | ···················· | ····················· | ···················· | ···················· | ···················· |
| S | 5005 | hCoV-19/England/ALDP-B459FA/2020\|EPI_ISL_656286\|20 | ···················· | ···················· | ····················· | ···················· | ···················· | ···················· |
| S | 5006 | hCoV-19/England/ALDP-B46416/2020\|EPI_ISL_656287\|20 | ···················· | ···················· | ····················· | ···················· | ···················· | ···················· |

| | | | | | | | |
|---|---|---|---|---|---|---|---|
| S | 5007 hCoV-19/England/ALDP-B46DA2/2020\|EPI_ISL_656288\|20 | ···················· | ···················· | ···················· | ···················· | ···················· | ···················· |
| S | 5008 hCoV-19/England/ALDP-B461E2/2020\|EPI_ISL_656290\|20 | ···················· | ···················· | ···················· | ···················· | ···················· | ···················· |
| S | 5009 hCoV-19/England/ALDP-B45FAD/2020\|EPI_ISL_656291\|20 | ···················· | ···················· | ···················· | ···················· | ···················· | ···················· |
| S | 5010 hCoV-19/England/ALDP-B45D67/2020\|EPI_ISL_656293\|20 | ···················· | ···················· | ···················· | ···················· | ···················· | ···················· |
| S | 5011 hCoV-19/England/ALDP-B45FE9/2020\|EPI_ISL_656295\|20 | ···················· | ···················· | ···················· | ···················· | ···················· | ···················· |
| S | 5012 hCoV-19/England/ALDP-B46C4B/2020\|EPI_ISL_656296\|20 | ···················· | ···················· | ···················· | ···················· | ···················· | ···················· |
| S | 5013 hCoV-19/England/ALDP-B45FCB/2020\|EPI_ISL_656297\|20 | ···················· | ·········T·········· | ···················· | ···················· | ···················· | ···················· |
| S | 5014 hCoV-19/England/ALDP-B4662F/2020\|EPI_ISL_656298\|20 | ···················· | ···················· | ···················· | ···················· | ···················· | ···················· |
| S | 5015 hCoV-19/England/ALDP-B46847/2020\|EPI_ISL_656299\|20 | ···················· | ···················· | ···················· | ···················· | ···················· | ···················· |
| S | 5016 hCoV-19/England/ALDP-B47293/2020\|EPI_ISL_656300\|20 | ···················· | ·A·················· | ···················· | ···················· | ···················· | ···················· |
| S | 5017 hCoV-19/England/ALDP-B45EBF/2020\|EPI_ISL_656301\|20 | ···················· | ···················· | ···················· | ···················· | ···················· | ···················· |
| S | 5018 hCoV-19/England/ALDP-B46E36/2020\|EPI_ISL_656302\|20 | ···················· | ···················· | ···················· | ···················· | ···················· | ···················· |
| S | 5019 hCoV-19/England/ALDP-B472DF/2020\|EPI_ISL_656303\|20 | ···················· | ···················· | ···················· | ···················· | ···················· | ···················· |
| S | 5020 hCoV-19/England/ALDP-B4605E/2020\|EPI_ISL_656304\|20 | ···················· | ···················· | ···················· | ···················· | ···················· | ···················· |
| S | 5021 hCoV-19/England/ALDP-B46100/2020\|EPI_ISL_656305\|20 | ···················· | ···················· | ···················· | ···················· | ···················· | ···················· |
| S | 5022 hCoV-19/England/ALDP-B45CE2/2020\|EPI_ISL_656306\|20 | ···················· | ···················· | ···················· | ···················· | ···················· | ···················· |
| S | 5023 hCoV-19/England/ALDP-B45F34/2020\|EPI_ISL_656307\|20 | ···················· | ···················· | ···················· | ···················· | ···················· | ···················· |
| S | 5024 hCoV-19/England/ALDP-B464CB/2020\|EPI_ISL_656308\|20 | ···················· | ···················· | ···················· | ···················· | ···················· | ···················· |
| S | 5025 hCoV-19/England/ALDP-B45972/2020\|EPI_ISL_656309\|20 | ···················· | ···················· | ···················· | ···················· | ···················· | ···················· |
| S | 5026 hCoV-19/England/ALDP-B467C2/2020\|EPI_ISL_656310\|20 | ···················· | ···················· | ···················· | ···················· | ···················· | ···················· |
| S | 5027 hCoV-19/England/ALDP-B45E19/2020\|EPI_ISL_656311\|20 | ···················· | ···················· | ···················· | ···················· | ···················· | ···················· |
| S | 5028 hCoV-19/England/ALDP-B462EF/2020\|EPI_ISL_656312\|20 | ···················· | ···················· | ···················· | ···················· | ···················· | ···················· |
| S | 5029 hCoV-19/England/ALDP-B46285/2020\|EPI_ISL_656313\|20 | ···················· | ···················· | ···················· | ···················· | ···················· | ···················· |
| S | 5030 hCoV-19/England/ALDP-B45DC1/2020\|EPI_ISL_656314\|20 | ···················· | ···················· | ···················· | ···················· | ···················· | ···················· |
| S | 5031 hCoV-19/England/ALDP-B46003/2020\|EPI_ISL_656315\|20 | ···················· | ···················· | ···················· | ···················· | ···················· | ···················· |
| S | 5032 hCoV-19/England/ALDP-B4663E/2020\|EPI_ISL_656316\|20 | ···················· | ···················· | ···················· | ···················· | ···················· | ···················· |
| S | 5033 hCoV-19/England/ALDP-B45AAC/2020\|EPI_ISL_656318\|20 | ···················· | ···················· | ···················· | ···················· | ···················· | ···················· |
| S | 5034 hCoV-19/England/ALDP-B46926/2020\|EPI_ISL_656319\|20 | ···················· | ···················· | ···················· | ···················· | ···················· | ···················· |
| S | 5035 hCoV-19/England/ALDP-B46021/2020\|EPI_ISL_656320\|20 | ···················· | ·········T·········· | ···················· | ···················· | ···················· | ···················· |
| S | 5036 hCoV-19/England/ALDP-B45B21/2020\|EPI_ISL_656322\|20 | ···················· | ···················· | ···················· | ···················· | ···················· | ···················· |
| S | 5037 hCoV-19/England/ALDP-B45D58/2020\|EPI_ISL_656324\|20 | ···················· | ···················· | ···················· | ···················· | ···················· | ···················· |
| S | 5038 hCoV-19/England/ALDP-B4674A/2020\|EPI_ISL_656325\|20 | ···················· | ···················· | ···················· | ···················· | ···················· | ···················· |
| S | 5039 hCoV-19/England/ALDP-B463CE/2020\|EPI_ISL_656327\|20 | ···················· | ···················· | ···················· | ···················· | ···················· | ···················· |
| S | 5040 hCoV-19/England/ALDP-B46B3F/2020\|EPI_ISL_656329\|20 | ···················· | ···················· | ···················· | ···················· | ···················· | ···················· |
| S | 5041 hCoV-19/England/ALDP-B45D85/2020\|EPI_ISL_656330\|20 | ···················· | ···················· | ···················· | ···················· | ···················· | ···················· |
| S | 5042 hCoV-19/England/ALDP-B45D2B/2020\|EPI_ISL_656331\|20 | ···················· | ···················· | ···················· | ···················· | ···················· | ···················· |
| S | 5043 hCoV-19/England/ALDP-B46874/2020\|EPI_ISL_656332\|20 | ···················· | ···················· | ···················· | ···················· | ···················· | ···················· |
| S | 5044 hCoV-19/England/ALDP-B468DE/2020\|EPI_ISL_656333\|20 | ···················· | ···················· | ···················· | ···················· | ···················· | ···················· |
| S | 5045 hCoV-19/England/ALDP-B466C5/2020\|EPI_ISL_656334\|20 | ···················· | ···················· | ···················· | ···················· | ···················· | ···················· |
| S | 5046 hCoV-19/England/ALDP-B45BE5/2020\|EPI_ISL_656335\|20 | ···················· | ···················· | ···················· | ···················· | ···················· | ···················· |
| S | 5047 hCoV-19/England/ALDP-B46768/2020\|EPI_ISL_656336\|20 | ···················· | ···················· | ···················· | ···················· | ···················· | ···················· |
| S | 5048 hCoV-19/England/ALDP-B460F4/2020\|EPI_ISL_656337\|20 | ···················· | ···················· | ···················· | ···················· | ···················· | ···················· |
| S | 5049 hCoV-19/England/ALDP-B4681A/2020\|EPI_ISL_656338\|20 | ···················· | ···················· | ···················· | ···················· | ···················· | ···················· |
| S | 5050 hCoV-19/England/CAMC-B52893/2020\|EPI_ISL_657590\|20 | ···················· | ···················· | ···················· | ···················· | ···················· | ···················· |
| S | 5051 hCoV-19/England/MILK-B555D5/2020\|EPI_ISL_657591\|20 | ···················· | ···················· | ···················· | ···················· | ···················· | ···················· |
| S | 5052 hCoV-19/England/CAMC-B53012/2020\|EPI_ISL_657592\|20 | ···················· | ···················· | ···················· | ···················· | ···················· | ···················· |
| S | 5053 hCoV-19/England/MILK-B555F3/2020\|EPI_ISL_657593\|20 | ···················· | ···················· | ···················· | ···················· | ···················· | ···A················ |
| S | 5054 hCoV-19/England/CAMC-B52BC7/2020\|EPI_ISL_657595\|20 | ···················· | ···················· | ···················· | ···················· | ···················· | ···················· |
| S | 5055 hCoV-19/England/MILK-B558AF/2020\|EPI_ISL_657596\|20 | ···················· | ·A·················· | ···················· | ···················· | ···················· | ···A················ |
| S | 5056 hCoV-19/England/CAMC-B52C88/2020\|EPI_ISL_657597\|20 | ···················· | ···················· | ···················· | ···················· | ···················· | ···················· |
| S | 5057 hCoV-19/England/CAMC-B52DEF/2020\|EPI_ISL_657598\|20 | ···················· | ···················· | ···················· | ···················· | ···················· | ···················· |
| S | 5058 hCoV-19/England/MILK-B567A1/2020\|EPI_ISL_657599\|20 | ···················· | ···················· | ···················· | ···················· | ···················· | ···················· |
| S | 5059 hCoV-19/England/CAMC-B52EA0/2020\|EPI_ISL_657626\|20 | ···················· | ···················· | ···················· | ···················· | ···················· | ···················· |
| S | 5060 hCoV-19/England/MILK-B55809/2020\|EPI_ISL_657627\|20 | ···················· | ···················· | ···················· | ···················· | ···················· | ···················· |
| S | 5061 hCoV-19/England/CAMC-B52E64/2020\|EPI_ISL_657628\|20 | ···················· | ···················· | ···················· | ···················· | ···················· | ···················· |
| S | 5062 hCoV-19/England/MILK-B566E0/2020\|EPI_ISL_657629\|20 | ···················· | ···················· | ···················· | ···················· | ···················· | ···················· |
| S | 5063 hCoV-19/England/MILK-B5646E/2020\|EPI_ISL_657631\|20 | ···················· | ···················· | ···················· | ···················· | ···················· | ···················· |
| S | 5064 hCoV-19/England/MILK-B562CE/2020\|EPI_ISL_657632\|20 | ···················· | ···················· | ···················· | ···················· | ···················· | ···················· |
| S | 5065 hCoV-19/England/CAMC-B53249/2020\|EPI_ISL_657633\|20 | ···················· | ···················· | ···················· | ···················· | ···················· | ···················· |
| S | 5066 hCoV-19/England/CAMC-B5315B/2020\|EPI_ISL_657634\|20 | ···················· | ···················· | ···················· | ···················· | ···················· | ···················· |
| S | 5067 hCoV-19/England/MILK-B5664A/2020\|EPI_ISL_657635\|20 | ···················· | ···················· | ···················· | ···················· | ···················· | ···················· |
| S | 5068 hCoV-19/England/MILK-B5648C/2020\|EPI_ISL_657636\|20 | ···················· | ···················· | ···················· | ···················· | ···················· | ···················· |
| S | 5069 hCoV-19/England/CAMC-B5281B/2020\|EPI_ISL_657637\|20 | ···················· | ···················· | ···················· | ···················· | ······T············· | ···················· |
| S | 5070 hCoV-19/England/MILK-B562DD/2020\|EPI_ISL_657639\|20 | ···················· | ···················· | ···················· | ···················· | ···················· | ···················· |
| S | 5071 hCoV-19/England/MILK-B56695/2020\|EPI_ISL_657640\|20 | ···················· | ·A·················· | ···················· | ···················· | ···················· | ···A················ |
| S | 5072 hCoV-19/England/MILK-B5620A/2020\|EPI_ISL_657642\|20 | ···················· | ···················· | ···················· | ···················· | ···················· | ···················· |
| S | 5073 hCoV-19/England/MILK-B556C3/2020\|EPI_ISL_657644\|20 | ···················· | ···················· | ···················· | ···················· | ···················· | ···················· |
| S | 5074 hCoV-19/England/CAMC-B5316A/2020\|EPI_ISL_657645\|20 | ···················· | ···················· | ···················· | ···················· | ···················· | ···················· |
| S | 5075 hCoV-19/England/MILK-B559D9/2020\|EPI_ISL_657647\|20 | ···················· | ···················· | ···················· | ···················· | ···················· | ···················· |
| S | 5076 hCoV-19/England/MILK-B55863/2020\|EPI_ISL_657648\|20 | ···················· | ···················· | ···················· | ···················· | ···················· | ···················· |
| S | 5077 hCoV-19/England/CAMC-B52FE9/2020\|EPI_ISL_657649\|20 | ···················· | ···················· | ···················· | ···················· | ···················· | ···················· |
| S | 5078 hCoV-19/England/MILK-B56422/2020\|EPI_ISL_657650\|20 | ···················· | ···················· | ···················· | ···················· | ···················· | ···A················ |
| S | 5079 hCoV-19/England/MILK-B55AD6/2020\|EPI_ISL_657651\|20 | ···················· | ···················· | ···················· | ···················· | ···················· | ···················· |
| S | 5080 hCoV-19/England/CAMC-B52F25/2020\|EPI_ISL_657656\|20 | ···················· | ···················· | ···················· | ···················· | ······T············· | ···················· |
| S | 5081 hCoV-19/England/MILK-B5656B/2020\|EPI_ISL_657657\|20 | ···················· | ···················· | ···················· | ···················· | ···················· | ···················· |
| S | 5082 hCoV-19/England/CAMC-B530F4/2020\|EPI_ISL_657658\|20 | ···················· | ···················· | ···················· | ···················· | ···················· | ···················· |
| S | 5083 hCoV-19/England/CAMC-B53319/2020\|EPI_ISL_657659\|20 | ···················· | ···················· | ···················· | ···················· | ···················· | ···················· |
| S | 5084 hCoV-19/England/CAMC-B52E19/2020\|EPI_ISL_657660\|20 | ···················· | ···················· | ···················· | ···················· | ···················· | ···················· |
| S | 5085 hCoV-19/England/MILK-B566B3/2020\|EPI_ISL_657662\|20 | ···················· | ···················· | ···················· | ···················· | ···················· | ···················· |
| S | 5086 hCoV-19/England/MILK-B56352/2020\|EPI_ISL_657663\|20 | ···················· | ···················· | ···················· | ···················· | ···················· | ···················· |

| | | | | | | | |
|---|---|---|---|---|---|---|---|
| S | 5087 hCoV-19/England/CAMC-B53258/2020\|EPI_ISL_657664\|20 | ···················· | ···················· | ····················· | ···················· | ···················· | ···················· |
| S | 5088 hCoV-19/England/MILK-B5572A/2020\|EPI_ISL_657665\|20 | ···················· | ···················· | ····················· | ···················· | ···················· | ···················· |
| S | 5089 hCoV-19/England/MILK-B55B1F/2020\|EPI_ISL_657666\|20 | ···················· | ·A·················· | ····················· | ···················· | ···················· | ···A················ |
| S | 5090 hCoV-19/England/CAMC-B52EEC/2020\|EPI_ISL_657668\|20 | ···················· | ···················· | ····················· | ···················· | ···················· | ···················· |
| S | 5091 hCoV-19/England/CAMC-B52AE8/2020\|EPI_ISL_657669\|20 | ···················· | ···················· | ····················· | ···················· | ···················· | ···················· |
| S | 5092 hCoV-19/England/MILK-B3A275/2020\|EPI_ISL_658957\|20 | ···················· | ···················· | ····················· | ···················· | ···················· | ···················· |
| S | 5093 hCoV-19/England/CAMC-B381C5/2020\|EPI_ISL_658959\|20 | ···················· | ···················· | ····················· | ···················· | ···················· | ···················· |
| S | 5094 hCoV-19/England/MILK-B38417/2020\|EPI_ISL_658962\|20 | ···················· | ···················· | ····················· | ···················· | ···················· | ···················· |
| S | 5095 hCoV-19/England/CAMC-B37EB0/2020\|EPI_ISL_658963\|20 | ···················· | ···················· | ····················· | ···················· | ···················· | ···················· |
| S | 5096 hCoV-19/England/MILK-B38B8B/2020\|EPI_ISL_658965\|20 | ···················· | ···················· | ····················· | ···················· | ···················· | ···················· |
| S | 5097 hCoV-19/England/MILK-B38D0D/2020\|EPI_ISL_658966\|20 | ···················· | ···················· | ····················· | ···················· | ···················· | ···················· |
| S | 5098 hCoV-19/England/MILK-B383B0/2020\|EPI_ISL_658967\|20 | ···················· | ···················· | ····················· | ···················· | ···················· | ···················· |
| S | 5099 hCoV-19/England/MILK-B38848/2020\|EPI_ISL_658968\|20 | ···················· | ···················· | ····················· | ···················· | ···················· | ···················· |
| S | 5100 hCoV-19/England/CAMC-B37FF9/2020\|EPI_ISL_658970\|20 | ···················· | ···················· | ····················· | ···················· | ···················· | ···················· |
| S | 5101 hCoV-19/England/MILK-B38C5B/2020\|EPI_ISL_658972\|20 | ···················· | ···················· | ····················· | ···················· | ···················· | ···················· |
| S | 5102 hCoV-19/England/MILK-B3856F/2020\|EPI_ISL_658973\|20 | ···················· | ···················· | ····················· | ···················· | ···················· | ···················· |
| S | 5103 hCoV-19/England/MILK-B38778/2020\|EPI_ISL_658974\|20 | ···················· | ·A·················· | ····················· | ···················· | ···················· | ···A················ |
| S | 5104 hCoV-19/England/MILK-B393CE/2020\|EPI_ISL_658975\|20 | ···················· | ···················· | ····················· | ···················· | ···················· | ···················· |
| S | 5105 hCoV-19/England/CAMC-B380C8/2020\|EPI_ISL_658976\|20 | ···················· | ···················· | ····················· | ···················· | ···················· | ···················· |
| S | 5106 hCoV-19/England/CAMC-B3821D/2020\|EPI_ISL_658979\|20 | ···················· | ···················· | ····················· | ···················· | ···················· | ···················· |
| S | 5107 hCoV-19/England/MILK-B38C88/2020\|EPI_ISL_658982\|20 | ···················· | ···················· | ····················· | ···················· | ···················· | ···················· |
| S | 5108 hCoV-19/England/MILK-B3870F/2020\|EPI_ISL_658983\|20 | ···················· | ···················· | ····················· | ···················· | ···················· | ···················· |
| S | 5109 hCoV-19/England/MILK-B39461/2020\|EPI_ISL_658984\|20 | ···················· | ···················· | ····················· | ···················· | ···················· | ···················· |
| S | 5110 hCoV-19/England/MILK-B3A5C7/2020\|EPI_ISL_658985\|20 | ···················· | ···················· | ····················· | ···················· | ···················· | ···················· |
| S | 5111 hCoV-19/England/MILK-B3921C/2020\|EPI_ISL_658987\|20 | ···················· | ···················· | ····················· | ···················· | ···················· | ···················· |
| S | 5112 hCoV-19/England/MILK-B38ABB/2020\|EPI_ISL_658988\|20 | ···················· | ···················· | ····················· | ···················· | ···················· | ···················· |
| S | 5113 hCoV-19/England/MILK-B38CB5/2020\|EPI_ISL_658989\|20 | ···················· | ···················· | ····················· | ···················· | ···················· | ···················· |
| S | 5114 hCoV-19/England/MILK-B38550/2020\|EPI_ISL_658990\|20 | ···················· | ·A·················· | ····················· | ···················· | ···················· | ···A················ |
| S | 5115 hCoV-19/England/MILK-B383A1/2020\|EPI_ISL_658991\|20 | ···················· | ···················· | ····················· | ···················· | ···················· | ···················· |
| S | 5116 hCoV-19/England/MILK-B38AAC/2020\|EPI_ISL_658992\|20 | ···················· | ···················· | ····················· | ···················· | ···················· | ···················· |
| S | 5117 hCoV-19/England/MILK-B38BC7/2020\|EPI_ISL_658993\|20 | ···················· | ···················· | ····················· | ···················· | ···················· | ···················· |
| S | 5118 hCoV-19/England/CAMC-B37D1D/2020\|EPI_ISL_658995\|20 | ···················· | ·A·················· | ····················· | ···················· | ···················· | ···A················ |
| S | 5119 hCoV-19/England/MILK-B38C97/2020\|EPI_ISL_658997\|20 | ···················· | ···················· | ····················· | ···················· | ···················· | ···················· |
| S | 5120 hCoV-19/England/MILK-B387D2/2020\|EPI_ISL_658999\|20 | ···················· | ·A·················· | ····················· | ···················· | ···················· | ···A················ |
| S | 5121 hCoV-19/New Zealand/20CV0550/2020\|EPI_ISL_649121\|2 | ···················· | ···················· | ····················· | ···················· | ···················· | ···················· |
| S | 5122 hCoV-19/New Zealand/20CV0554/2020\|EPI_ISL_649122\|2 | ···················· | ·A·················· | ····················· | ···················· | ···················· | ···················· |
| S | 5123 hCoV-19/New Zealand/20CV0556/2020\|EPI_ISL_649123\|2 | ···················· | ·A·················· | ····················· | ···················· | ···················· | ···A················ |
| S | 5124 hCoV-19/New Zealand/20CV0558/2020\|EPI_ISL_649124\|2 | ···················· | ···················· | ····················· | ···················· | ···················· | ···················· |
| S | 5125 hCoV-19/New Zealand/20CV0563/2020\|EPI_ISL_649125\|2 | ···················· | ·A·················· | ····················· | ···················· | ···················· | ···A················ |
| S | 5129 hCoV-19/USA/MI-UM-10036634217/2020\|EPI_ISL_650040\| | ···················· | ···················· | ····················· | ···················· | ···················· | ···················· |
| S | 5131 hCoV-19/USA/MI-UM-10036624405/2020\|EPI_ISL_650043\| | ···················· | ·············A······ | ····················· | ···················· | ···················· | ···················· |
| S | 5132 hCoV-19/USA/MI-UM-10036633055/2020\|EPI_ISL_650044\| | ···················· | ·A·················· | ····················· | ···················· | ···················· | ···················· |
| S | 5133 hCoV-19/USA/MI-UM-10036633033/2020\|EPI_ISL_650045\| | ···················· | ···················· | ····················· | ···················· | ···················· | ···················· |
| S | 5136 hCoV-19/USA/MI-UM-10036643724/2020\|EPI_ISL_650048\| | ···················· | ···················· | ····················· | ···················· | ···················· | ···················· |
| S | 5137 hCoV-19/USA/MI-UM-10036661466/2020\|EPI_ISL_650049\| | ···················· | ···················· | ····················· | ···················· | ···················· | ···················· |
| S | 5139 hCoV-19/USA/MI-UM-10036648902/2020\|EPI_ISL_650051\| | ···················· | ···················· | ····················· | ···················· | ···················· | ···················· |
| S | 5141 hCoV-19/USA/MI-UM-10036671739/2020\|EPI_ISL_650053\| | ···················· | ···················· | ····················· | ···················· | ···················· | ···················· |
| S | 5142 hCoV-19/USA/MI-UM-10036667320/2020\|EPI_ISL_650054\| | ···················· | ···················· | ····················· | ···················· | ···················· | ···················· |
| S | 5143 hCoV-19/USA/MI-UM-10036629769/2020\|EPI_ISL_650055\| | ···················· | ···················· | ····················· | ···················· | ···················· | ···················· |
| S | 5144 hCoV-19/USA/MI-UM-10036672505/2020\|EPI_ISL_650056\| | ···················· | ···················· | ····················· | ···················· | ···················· | ···················· |
| S | 5145 hCoV-19/USA/MI-UM-10036665971/2020\|EPI_ISL_650057\| | ···················· | ···················· | ····················· | ···················· | ···················· | ···················· |
| S | 5146 hCoV-19/USA/MI-UM-10036638082/2020\|EPI_ISL_650058\| | ···················· | ···················· | ····················· | ···················· | ···················· | ···················· |
| S | 5147 hCoV-19/USA/MI-UM-10036648807/2020\|EPI_ISL_650059\| | ···················· | ···················· | ····················· | ···················· | ···················· | ···················· |
| S | 5148 hCoV-19/USA/MI-UM-10036629202/2020\|EPI_ISL_650060\| | ···················· | ···················· | ····················· | ···················· | ···················· | ···················· |
| S | 5149 hCoV-19/USA/MI-UM-10036664297/2020\|EPI_ISL_650063\| | ···················· | ···················· | ····················· | ···················· | ···················· | ···················· |
| S | 5150 hCoV-19/USA/MI-UM-10036641218/2020\|EPI_ISL_650064\| | ···················· | ···················· | ····················· | ···················· | ···················· | ···················· |
| S | 5152 hCoV-19/USA/MI-UM-10036681654/2020\|EPI_ISL_650066\| | ···················· | ···················· | ····················· | ···················· | ···················· | ···················· |
| S | 5155 hCoV-19/USA/MI-UM-10036652026/2020\|EPI_ISL_650069\| | ···················· | ···················· | ····················· | ···················· | ···················· | ···················· |
| S | 5160 hCoV-19/USA/MI-UM-10036648750/2020\|EPI_ISL_650077\| | ···················· | ···················· | ····················· | ···················· | ···················· | ···················· |
| S | 5171 hCoV-19/USA/MI-UM-10036671636/2020\|EPI_ISL_650093\| | ···················· | ·A·················· | ······G·············· | ···················· | ···················· | ···················· |
| S | 5175 hCoV-19/USA/MI-UM-10036669304/2020\|EPI_ISL_650097\| | ···················· | ···················· | ····················· | ···················· | ···················· | ···················· |
| S | 5177 hCoV-19/USA/MI-UM-10036687960/2020\|EPI_ISL_650099\| | ···················· | ···················· | ····················· | ···················· | ···················· | ···················· |
| S | 5179 hCoV-19/USA/MI-UM-10036665972/2020\|EPI_ISL_650101\| | ···················· | ···················· | ····················· | ···················· | ···················· | ···················· |
| S | 5184 hCoV-19/England/CAMC-B23127/2020\|EPI_ISL_651414\|20 | ···················· | ···················· | ····················· | ···················· | ···················· | ···················· |
| S | 5185 hCoV-19/Wales/PHWC-489E01/2020\|EPI_ISL_651415\|2020 | ···················· | ···················· | ····················· | ···················· | ···················· | ···················· |
| S | 5186 hCoV-19/Wales/PHWC-48A36B/2020\|EPI_ISL_651416\|2020 | ···················· | ···················· | ····················· | ···················· | ···················· | ···················· |
| S | 5187 hCoV-19/England/QEUH-AACA59/2020\|EPI_ISL_651417\|20 | ···················· | ···················· | ····················· | ···················· | ···················· | ···················· |
| S | 5188 hCoV-19/England/QEUH-AF5FF8/2020\|EPI_ISL_651418\|20 | ···················· | ···················· | ····················· | ···················· | ···················· | ···················· |
| S | 5189 hCoV-19/England/QEUH-AF60A9/2020\|EPI_ISL_651419\|20 | ···················· | ···················· | ····················· | ···················· | ···················· | ···················· |
| S | 5190 hCoV-19/Wales/PHWC-48986D/2020\|EPI_ISL_651420\|2020 | ···················· | ···················· | ····················· | ···················· | ···················· | ···················· |
| S | 5191 hCoV-19/England/QEUH-AAB270/2020\|EPI_ISL_651421\|20 | ···················· | ···················· | ····················· | ···················· | ···················· | ···················· |
| S | 5192 hCoV-19/England/BIRM-6ABA9/2020\|EPI_ISL_651422\|202 | ···················· | ···················· | ····················· | ···················· | ···················· | ···················· |
| S | 5193 hCoV-19/England/CAMB-1B88C2/2020\|EPI_ISL_651423\|20 | ···················· | ···················· | ····················· | ···················· | ···················· | ···················· |
| S | 5194 hCoV-19/Scotland/CVR5417/2020\|EPI_ISL_651424\|2020- | ···················· | ···················· | ····················· | ···················· | ···················· | ···················· |
| S | 5195 hCoV-19/Scotland/EDB8808/2020\|EPI_ISL_651425\|2020- | ···················· | ···················· | ····················· | ···················· | ···················· | ···················· |
| S | 5196 hCoV-19/Scotland/EDB8819/2020\|EPI_ISL_651426\|2020- | ···················· | ···················· | ····················· | ···················· | ···················· | ···················· |
| S | 5197 hCoV-19/Scotland/EDB8974/2020\|EPI_ISL_651427\|2020- | ···················· | ···················· | ····················· | ···················· | ···················· | ···················· |
| S | 5198 hCoV-19/Scotland/EDB9228/2020\|EPI_ISL_651428\|2020- | ···················· | ···················· | ····················· | ···················· | ···················· | ···················· |
| S | 5199 hCoV-19/England/LIVE-DB4B0D/2020\|EPI_ISL_651429\|20 | ···················· | ···················· | ····················· | ···················· | ···················· | ···················· |
| S | 5200 hCoV-19/England/LIVE-DB64A6/2020\|EPI_ISL_651430\|20 | ···················· | ···················· | ····················· | ···················· | ···················· | ···················· |

| | | | | | | | |
|---|---|---|---|---|---|---|---|
| S | 5201 hCoV-19/England/MILK-ACFEAF/2020\|EPI_ISL_651433\|20 | ···················· | ···················· | ···················· | ···················· | ···················· | ···················· |
| S | 5202 hCoV-19/England/MILK-B02FCA/2020\|EPI_ISL_651434\|20 | ···················· | ···················· | ···················· | ···················· | ···················· | ···················· |
| S | 5203 hCoV-19/England/MILK-B0461E/2020\|EPI_ISL_651435\|20 | ···················· | ···················· | ···················· | ···················· | ···················· | ···················· |
| S | 5204 hCoV-19/Wales/MILK-B04E25/2020\|EPI_ISL_651436\|2020 | ···················· | ···················· | ···················· | ···················· | ···················· | ···················· |
| S | 5205 hCoV-19/Wales/MILK-B05413/2020\|EPI_ISL_651437\|2020 | ···················· | ···················· | ···················· | ···················· | ···················· | ···················· |
| S | 5206 hCoV-19/England/MILK-B06676/2020\|EPI_ISL_651438\|20 | ···················· | ···················· | ···················· | ···················· | ···················· | ···················· |
| S | 5207 hCoV-19/England/MILK-B392C1/2020\|EPI_ISL_651439\|20 | ···················· | ···················· | ···················· | ···················· | ···················· | ···················· |
| S | 5208 hCoV-19/Wales/PHWC-48984F/2020\|EPI_ISL_651440\|2020 | ···················· | ···················· | ···················· | ···················· | ···················· | ···················· |
| S | 5209 hCoV-19/Wales/PHWC-4898F4/2020\|EPI_ISL_651441\|2020 | ···················· | ···················· | ···················· | ···················· | ···················· | ···················· |
| S | 5210 hCoV-19/Wales/PHWC-489979/2020\|EPI_ISL_651442\|2020 | ···················· | ···················· | ···················· | ···················· | ···················· | ···················· |
| S | 5211 hCoV-19/Wales/PHWC-4899F1/2020\|EPI_ISL_651443\|2020 | ···················· | ···················· | ···················· | ···················· | ···················· | ···················· |
| S | 5212 hCoV-19/England/QEUH-A9FFC3/2020\|EPI_ISL_652698\|20 | ···················· | ···················· | ···················· | ···················· | ···················· | ···················· |
| S | 5213 hCoV-19/England/QEUH-A9FFF0/2020\|EPI_ISL_652699\|20 | ···················· | ···················· | ···················· | ···················· | ···················· | ···················· |
| S | 5214 hCoV-19/Scotland/QEUH-AAAAA6/2020\|EPI_ISL_652731\|2 | ···················· | ···················· | ···················· | ···················· | ···················· | ···················· |
| S | 5215 hCoV-19/England/QEUH-AAABEF/2020\|EPI_ISL_652732\|20 | ···················· | ···················· | ···················· | ···················· | ···················· | ···················· |
| S | 5216 hCoV-19/England/QEUH-AAABFE/2020\|EPI_ISL_652733\|20 | ···················· | ···················· | ···················· | ···················· | ···················· | ···················· |
| S | 5217 hCoV-19/England/QEUH-AAAC37/2020\|EPI_ISL_652734\|20 | ···················· | ···················· | ···················· | ···················· | ···················· | ···················· |
| S | 5218 hCoV-19/England/QEUH-AAAC55/2020\|EPI_ISL_652735\|20 | ···················· | ···················· | ···················· | ···················· | ···················· | ···················· |
| S | 5219 hCoV-19/England/QEUH-AAAC64/2020\|EPI_ISL_652736\|20 | ···················· | ·A·················· | ···················· | ···················· | ···················· | ··A················· |
| S | 5220 hCoV-19/England/QEUH-AAAC73/2020\|EPI_ISL_652737\|20 | ···················· | ···················· | ···················· | ···················· | ···················· | ···················· |
| S | 5221 hCoV-19/England/QEUH-AAAC82/2020\|EPI_ISL_652738\|20 | ···················· | ···················· | ···················· | ···················· | ···················· | ···················· |
| S | 5222 hCoV-19/England/QEUH-AAAC91/2020\|EPI_ISL_652739\|20 | ···················· | ·A·················· | ···················· | ···················· | ···················· | ··A················· |
| S | 5223 hCoV-19/England/QEUH-AAACA0/2020\|EPI_ISL_652740\|20 | ···················· | ···················· | ···················· | ···················· | ···················· | ···················· |
| S | 5224 hCoV-19/England/QEUH-AAAD9E/2020\|EPI_ISL_652741\|20 | ···················· | ···················· | ···················· | ···················· | ···················· | ···················· |
| S | 5225 hCoV-19/England/QEUH-AAADAD/2020\|EPI_ISL_652742\|20 | ···················· | ···················· | ···················· | ···················· | ···················· | ···················· |
| S | 5226 hCoV-19/England/QEUH-AAADDA/2020\|EPI_ISL_652743\|20 | ···················· | ···················· | ···················· | ···················· | ···················· | ···················· |
| S | 5227 hCoV-19/England/QEUH-AAADE9/2020\|EPI_ISL_652744\|20 | ···················· | ···················· | ···················· | ···················· | ···················· | ···················· |
| S | 5228 hCoV-19/England/QEUH-AAAE04/2020\|EPI_ISL_652745\|20 | ···················· | ···················· | ···················· | ···················· | ···················· | ···················· |
| S | 5229 hCoV-19/England/QEUH-AAAE13/2020\|EPI_ISL_652746\|20 | ···················· | ···················· | ···················· | ···················· | ···················· | ···················· |
| S | 5230 hCoV-19/England/QEUH-AAAE22/2020\|EPI_ISL_652747\|20 | ···················· | ···················· | ···················· | ···················· | ···················· | ···················· |
| S | 5231 hCoV-19/England/QEUH-AAAE31/2020\|EPI_ISL_652748\|20 | ···················· | ···················· | ···················· | ···················· | ···················· | ···················· |
| S | 5232 hCoV-19/England/QEUH-AAAE5F/2020\|EPI_ISL_652749\|20 | ···················· | ···················· | ···················· | ···················· | ···················· | ···················· |
| S | 5233 hCoV-19/England/QEUH-AAAE6E/2020\|EPI_ISL_652750\|20 | ···················· | ···················· | ···················· | ···················· | ···················· | ···················· |
| S | 5234 hCoV-19/England/QEUH-AAAE9B/2020\|EPI_ISL_652751\|20 | ···················· | ···················· | ···················· | ···················· | ···················· | ···················· |
| S | 5235 hCoV-19/England/QEUH-AAAEC8/2020\|EPI_ISL_652752\|20 | ···················· | ···················· | ···················· | ···················· | ···················· | ···················· |
| S | 5236 hCoV-19/England/QEUH-AAAED7/2020\|EPI_ISL_652753\|20 | ···················· | ···················· | ···················· | ···················· | ······T············· | ···················· |
| S | 5237 hCoV-19/England/QEUH-AAAEF5/2020\|EPI_ISL_652754\|20 | ···················· | ···················· | ···················· | ···················· | ···················· | ···················· |
| S | 5238 hCoV-19/England/QEUH-AAAF10/2020\|EPI_ISL_652755\|20 | ···················· | ···················· | ···················· | ···················· | ···················· | ···················· |
| S | 5239 hCoV-19/England/QEUH-AAAF4D/2020\|EPI_ISL_652756\|20 | ···················· | ···················· | ···················· | ···················· | ···················· | ···················· |
| S | 5240 hCoV-19/England/QEUH-AAAF5C/2020\|EPI_ISL_652757\|20 | ···················· | ···················· | ···················· | ···················· | ···················· | ···················· |
| S | 5241 hCoV-19/England/QEUH-AAAF6B/2020\|EPI_ISL_652758\|20 | ···················· | ···················· | ···················· | ···················· | ···················· | ···················· |
| S | 5242 hCoV-19/England/QEUH-AAAF7A/2020\|EPI_ISL_652759\|20 | ···················· | ···················· | ···················· | ···················· | ···················· | ···················· |
| S | 5243 hCoV-19/England/QEUH-AAAF98/2020\|EPI_ISL_652761\|20 | ···················· | ···················· | ···················· | ···················· | ···················· | ···················· |
| S | 5244 hCoV-19/England/QEUH-AAAFE3/2020\|EPI_ISL_652764\|20 | ···················· | ···················· | ···················· | ···················· | ···················· | ···················· |
| S | 5245 hCoV-19/England/QEUH-AAB00D/2020\|EPI_ISL_652765\|20 | ···················· | ···················· | ···················· | ···················· | ···················· | ···················· |
| S | 5246 hCoV-19/England/QEUH-AAB01C/2020\|EPI_ISL_652766\|20 | ···················· | ···················· | ···················· | ···················· | ···················· | ···················· |
| S | 5247 hCoV-19/England/QEUH-AAB03A/2020\|EPI_ISL_652767\|20 | ···················· | ···················· | ···················· | ···················· | ···················· | ···················· |
| S | 5248 hCoV-19/England/QEUH-AAB076/2020\|EPI_ISL_652768\|20 | ···················· | ···················· | ···················· | ···················· | ···················· | ···················· |
| S | 5249 hCoV-19/England/QEUH-AAB0B2/2020\|EPI_ISL_652769\|20 | ···················· | ···················· | ···················· | ···················· | ···················· | ···················· |
| S | 5250 hCoV-19/England/QEUH-AAB0C1/2020\|EPI_ISL_652770\|20 | ···················· | ·A·················· | ···················· | ···················· | ···················· | ··A················· |
| S | 5251 hCoV-19/England/QEUH-AAB0D0/2020\|EPI_ISL_652771\|20 | ···················· | ···················· | ···················· | ···················· | ···················· | ···················· |
| S | 5252 hCoV-19/England/QEUH-AAB0FE/2020\|EPI_ISL_652772\|20 | ···················· | ···················· | ···················· | ···················· | ···················· | ···················· |
| S | 5253 hCoV-19/England/QEUH-AAB10A/2020\|EPI_ISL_652773\|20 | ···················· | ···················· | ···················· | ···················· | ···················· | ···················· |
| S | 5254 hCoV-19/Scotland/QEUH-AAB119/2020\|EPI_ISL_652774\|2 | ···················· | ···················· | ···················· | ···················· | ···················· | ···················· |
| S | 5255 hCoV-19/England/ALDP-B46BC6/2020\|EPI_ISL_656218\|20 | ···················· | ···················· | ···················· | ···················· | ···················· | ···················· |
| S | 5256 hCoV-19/England/ALDP-B468B0/2020\|EPI_ISL_656220\|20 | ···················· | ···················· | ···················· | ···················· | ···················· | ···················· |
| S | 5257 hCoV-19/England/ALDP-B45DD0/2020\|EPI_ISL_656222\|20 | ···················· | ···················· | ···················· | ···················· | ···················· | ···················· |
| S | 5258 hCoV-19/England/ALDP-B461C4/2020\|EPI_ISL_656223\|20 | ···················· | ···················· | ···················· | ···················· | ···················· | ···················· |
| S | 5259 hCoV-19/England/ALDP-B46CC3/2020\|EPI_ISL_656224\|20 | ···················· | ···················· | ···················· | ···················· | ···················· | ···················· |
| S | 5260 hCoV-19/England/ALDP-B469AE/2020\|EPI_ISL_656225\|20 | ···················· | ···················· | ···················· | ···················· | ···················· | ···················· |
| S | 5261 hCoV-19/England/ALDP-B45D3A/2020\|EPI_ISL_656226\|20 | ···················· | ···················· | ···················· | ···················· | ···················· | ···················· |
| S | 5262 hCoV-19/England/ALDP-B46BA8/2020\|EPI_ISL_656227\|20 | ···················· | ···················· | ···················· | ···················· | ···················· | ···················· |
| S | 5263 hCoV-19/England/ALDP-B46A7E/2020\|EPI_ISL_656228\|20 | ···················· | ···················· | ···················· | ···················· | ···················· | ···················· |
| S | 5264 hCoV-19/England/ALDP-B46D48/2020\|EPI_ISL_656229\|20 | ···················· | ···················· | ···················· | ···················· | ···················· | ···················· |
| S | 5265 hCoV-19/England/ALDP-B46E45/2020\|EPI_ISL_656231\|20 | ···················· | ···················· | ···················· | ···················· | ···················· | ···················· |
| S | 5266 hCoV-19/England/ALDP-B46D93/2020\|EPI_ISL_656232\|20 | ···················· | ···················· | ···················· | ···················· | ···················· | ···················· |
| S | 5267 hCoV-19/England/ALDP-B45DA3/2020\|EPI_ISL_656234\|20 | ···················· | ···················· | ···················· | ···················· | ···················· | ···················· |
| S | 5268 hCoV-19/England/ALDP-B464E9/2020\|EPI_ISL_656235\|20 | ···················· | ···················· | ···················· | ···················· | ···················· | ···················· |
| S | 5269 hCoV-19/England/ALDP-B46A50/2020\|EPI_ISL_656236\|20 | ···················· | ···················· | ···················· | ···················· | ···················· | ···················· |
| S | 5270 hCoV-19/England/ALDP-B4657D/2020\|EPI_ISL_656237\|20 | ···················· | ···················· | ···················· | ···················· | ···················· | ···················· |
| S | 5271 hCoV-19/England/ALDP-B46DDF/2020\|EPI_ISL_656238\|20 | ···················· | ···················· | ···················· | ···················· | ···················· | ···················· |
| S | 5272 hCoV-19/England/ALDP-B45C1F/2020\|EPI_ISL_656240\|20 | ···················· | ···················· | ···················· | ···················· | ···················· | ···················· |
| S | 5273 hCoV-19/England/ALDP-B4615B/2020\|EPI_ISL_656241\|20 | ···················· | ···················· | ···················· | ···················· | ···················· | ···················· |
| S | 5274 hCoV-19/England/ALDP-B465F5/2020\|EPI_ISL_656242\|20 | ···················· | ···················· | ···················· | ···················· | ···················· | ···················· |
| S | 5275 hCoV-19/England/ALDP-B465E6/2020\|EPI_ISL_656244\|20 | ···················· | ···················· | ···················· | ···················· | ···················· | ···················· |
| S | 5276 hCoV-19/England/ALDP-B466F2/2020\|EPI_ISL_656245\|20 | ···················· | ···················· | ···················· | ···················· | ···················· | ···················· |
| S | 5277 hCoV-19/England/ALDP-B45B12/2020\|EPI_ISL_656246\|20 | ···················· | ···················· | ···················· | ···················· | ···················· | ···················· |
| S | 5278 hCoV-19/England/ALDP-B46382/2020\|EPI_ISL_656247\|20 | ···················· | ·A·················· | ···················· | ···················· | ···················· | ··A················· |
| S | 5279 hCoV-19/England/ALDP-B464F8/2020\|EPI_ISL_656248\|20 | ···················· | ···················· | ···················· | ···················· | ···················· | ···················· |
| S | 5280 hCoV-19/England/ALDP-B46CD2/2020\|EPI_ISL_656249\|20 | ···················· | ···················· | ···················· | ···················· | ···················· | ···················· |

| | | | | | | | |
|---|---|---|---|---|---|---|---|
| S | 5281 hCoV-19/England/ALDP-B45F8F/2020\|EPI_ISL_656250\|20 | ···················· | ···················· | ···················· | ···················· | ···················· | ···················· |
| S | 5282 hCoV-19/England/ALDP-B46962/2020\|EPI_ISL_656251\|20 | ···················· | ···················· | ···················· | ···················· | ···················· | ···················· |
| S | 5283 hCoV-19/England/ALDP-B46795/2020\|EPI_ISL_656252\|20 | ···················· | ···················· | ···················· | ···················· | ···················· | ···················· |
| S | 5284 hCoV-19/England/ALDP-B46F06/2020\|EPI_ISL_656253\|20 | ···················· | ···················· | ···················· | ···················· | ···················· | ···················· |
| S | 5285 hCoV-19/England/ALDP-B45F70/2020\|EPI_ISL_656254\|20 | ···················· | ···················· | ···················· | ···················· | ···················· | ···················· |
| S | 5286 hCoV-19/England/ALDP-B45E73/2020\|EPI_ISL_656256\|20 | ···················· | ···················· | ···················· | ···················· | ···················· | ···················· |
| S | 5287 hCoV-19/England/ALDP-B4665C/2020\|EPI_ISL_656258\|20 | ···················· | ···················· | ···················· | ···················· | ···················· | ···················· |
| S | 5288 hCoV-19/England/ALDP-B46FE8/2020\|EPI_ISL_656259\|20 | ···················· | ···················· | ···················· | ···················· | ···················· | ···················· |
| S | 5289 hCoV-19/England/MILK-B56598/2020\|EPI_ISL_657500\|20 | ···················· | ···················· | ···················· | ···················· | ···················· | ···················· |
| S | 5290 hCoV-19/England/CAMC-B52E37/2020\|EPI_ISL_657501\|20 | ···················· | ···················· | ···················· | ···················· | ···················· | ···················· |
| S | 5291 hCoV-19/England/MILK-B56228/2020\|EPI_ISL_657503\|20 | ···················· | ···················· | ···················· | ···················· | ···················· | ···················· |
| S | 5292 hCoV-19/England/MILK-B56316/2020\|EPI_ISL_657550\|20 | ···················· | ·A·················· | ···················· | ···················· | ···················· | ··A················· |
| S | 5293 hCoV-19/England/CAMC-B52B21/2020\|EPI_ISL_657552\|20 | ···················· | ···················· | ···················· | ···················· | ······T············· | ···················· |
| S | 5294 hCoV-19/England/MILK-B562BF/2020\|EPI_ISL_657553\|20 | ···················· | ···················· | ···················· | ···················· | ···················· | ···················· |
| S | 5295 hCoV-19/England/MILK-B558BE/2020\|EPI_ISL_657554\|20 | ···················· | ···················· | ···················· | ···················· | ···················· | ···················· |
| S | 5296 hCoV-19/England/MILK-B557B1/2020\|EPI_ISL_657555\|20 | ···················· | ···················· | ···················· | ···················· | ···················· | ···················· |
| S | 5297 hCoV-19/England/MILK-B56370/2020\|EPI_ISL_657556\|20 | ···················· | ···················· | ···················· | ···················· | ···················· | ···················· |
| S | 5298 hCoV-19/England/MILK-B55951/2020\|EPI_ISL_657557\|20 | ···················· | ···················· | ···················· | ···················· | ···················· | ···················· |
| S | 5299 hCoV-19/England/MILK-B55748/2020\|EPI_ISL_657558\|20 | ···················· | ···················· | ···················· | ···················· | ···················· | ···················· |
| S | 5300 hCoV-19/England/CAMC-B53021/2020\|EPI_ISL_657559\|20 | ···················· | ···················· | ···················· | ···················· | ···················· | ···················· |
| S | 5301 hCoV-19/England/CAMC-B52C5B/2020\|EPI_ISL_657561\|20 | ···················· | ···················· | ···················· | ···················· | ···················· | ···················· |
| S | 5302 hCoV-19/England/CAMC-B52DD0/2020\|EPI_ISL_657562\|20 | ···················· | ···················· | ···················· | ···················· | ···················· | ···················· |
| S | 5303 hCoV-19/England/MILK-B559AC/2020\|EPI_ISL_657563\|20 | ···················· | ···················· | ···················· | ···················· | ···················· | ···················· |
| S | 5304 hCoV-19/England/MILK-B55A7C/2020\|EPI_ISL_657564\|20 | ···················· | ···················· | ···················· | ···················· | ···················· | ···················· |
| S | 5305 hCoV-19/England/CAMC-B52A51/2020\|EPI_ISL_657565\|20 | ···················· | ···················· | ···················· | ···················· | ···················· | ···················· |
| S | 5306 hCoV-19/England/CAMC-B53030/2020\|EPI_ISL_657566\|20 | ···················· | ···················· | ···················· | ···················· | ···················· | ···················· |
| S | 5307 hCoV-19/England/MILK-B5638F/2020\|EPI_ISL_657567\|20 | ···················· | ···················· | ···················· | ···················· | ···················· | ···················· |
| S | 5308 hCoV-19/England/MILK-B5561E/2020\|EPI_ISL_657568\|20 | ···················· | ···················· | ···················· | ···················· | ···················· | ···················· |
| S | 5309 hCoV-19/England/CAMC-B53003/2020\|EPI_ISL_657569\|20 | ···················· | ···················· | ···················· | ···················· | ···················· | ···················· |
| S | 5310 hCoV-19/England/MILK-B563CB/2020\|EPI_ISL_657570\|20 | ···················· | ···················· | ···················· | ···················· | ···················· | ···················· |
| S | 5311 hCoV-19/England/MILK-B5563C/2020\|EPI_ISL_657571\|20 | ···················· | ···················· | ···················· | ···················· | ···················· | ···················· |
| S | 5312 hCoV-19/England/CAMC-B52E46/2020\|EPI_ISL_657572\|20 | ···················· | ···················· | ···················· | ···················· | ···················· | ···················· |
| S | 5313 hCoV-19/England/CAMC-B52EFB/2020\|EPI_ISL_657573\|20 | ···················· | ···················· | ···················· | ···················· | ···················· | ···················· |
| S | 5314 hCoV-19/England/MILK-B5654D/2020\|EPI_ISL_657574\|20 | ···················· | ·A·················· | ···················· | ···················· | ···················· | ··A················· |
| S | 5315 hCoV-19/England/MILK-B56729/2020\|EPI_ISL_657575\|20 | ···················· | ···················· | ···················· | ···················· | ···················· | ···················· |
| S | 5316 hCoV-19/England/CAMC-B529EB/2020\|EPI_ISL_657576\|20 | ···················· | ···················· | ···················· | ···················· | ···················· | ···················· |
| S | 5317 hCoV-19/England/MILK-B55818/2020\|EPI_ISL_657577\|20 | ···················· | ···················· | ···················· | ···················· | ···················· | ···················· |
| S | 5318 hCoV-19/England/CAMC-B5282A/2020\|EPI_ISL_657578\|20 | ···················· | ···················· | ···················· | ···················· | ······T············· | ···················· |
| S | 5319 hCoV-19/England/MILK-B55775/2020\|EPI_ISL_657581\|20 | ···················· | ···················· | ···················· | ···················· | ···················· | ···················· |
| S | 5320 hCoV-19/England/CAMC-B52B12/2020\|EPI_ISL_657582\|20 | ···················· | ···················· | ···················· | ···················· | ···················· | ···················· |
| S | 5321 hCoV-19/England/MILK-B55A8B/2020\|EPI_ISL_657583\|20 | ···················· | ···················· | ···················· | ···················· | ···················· | ···················· |
| S | 5322 hCoV-19/England/MILK-B5560F/2020\|EPI_ISL_657584\|20 | ···················· | ···················· | ···················· | ···················· | ···················· | ···················· |
| S | 5323 hCoV-19/England/MILK-B55B3D/2020\|EPI_ISL_657585\|20 | ···················· | ···················· | ···················· | ···················· | ···················· | ···················· |
| S | 5324 hCoV-19/England/CAMC-B52F8F/2020\|EPI_ISL_657586\|20 | ···················· | ···················· | ···················· | ···················· | ···················· | ···················· |
| S | 5325 hCoV-19/England/MILK-B56501/2020\|EPI_ISL_657587\|20 | ···················· | ···················· | ···················· | ···················· | ···················· | ···················· |
| S | 5326 hCoV-19/England/CAMC-B527A5/2020\|EPI_ISL_657588\|20 | ···················· | ···················· | ···················· | ···················· | ······T············· | ···················· |
| S | 5327 hCoV-19/England/MILK-B56738/2020\|EPI_ISL_657589\|20 | ···················· | ···················· | ···················· | ···················· | ···················· | ···················· |
| S | 5328 hCoV-19/England/MILK-B3868A/2020\|EPI_ISL_658801\|20 | ···················· | ···················· | ···················· | ···················· | ···················· | ···················· |
| S | 5329 hCoV-19/England/MILK-B38C1F/2020\|EPI_ISL_658802\|20 | ···················· | ···················· | ···················· | ···················· | ···················· | ···················· |
| S | 5330 hCoV-19/England/MILK-B3A5E5/2020\|EPI_ISL_658803\|20 | ···················· | ···················· | ···················· | ···················· | ···················· | ···················· |
| S | 5331 hCoV-19/England/MILK-B385C9/2020\|EPI_ISL_658806\|20 | ···················· | ·A·················· | ···················· | ···················· | ···················· | ··A················· |
| S | 5332 hCoV-19/England/MILK-B38DFE/2020\|EPI_ISL_658808\|20 | ···················· | ···················· | ···················· | ···················· | ···················· | ···················· |
| S | 5333 hCoV-19/England/MILK-B38374/2020\|EPI_ISL_658810\|20 | ···················· | ···················· | ···················· | ···················· | ···················· | ···················· |
| S | 5334 hCoV-19/England/MILK-B38A15/2020\|EPI_ISL_658811\|20 | ···················· | ···················· | ···················· | ···················· | ···················· | ···················· |
| S | 5335 hCoV-19/England/MILK-B39522/2020\|EPI_ISL_658812\|20 | ···················· | ···················· | ···················· | ···················· | ···················· | ···················· |
| S | 5336 hCoV-19/England/MILK-B388A2/2020\|EPI_ISL_658813\|20 | ···················· | ···················· | ···················· | ···················· | ···················· | ···················· |
| S | 5337 hCoV-19/England/MILK-B38884/2020\|EPI_ISL_658814\|20 | ···················· | ···················· | ···················· | ···················· | ···················· | ···················· |
| S | 5338 hCoV-19/England/MILK-B393EC/2020\|EPI_ISL_658815\|20 | ···················· | ···················· | ···················· | ···················· | ···················· | ···················· |
| S | 5339 hCoV-19/England/CAMC-B37DFF/2020\|EPI_ISL_658816\|20 | ···················· | ···················· | ···················· | ···················· | ······T············· | ···················· |
| S | 5340 hCoV-19/England/MILK-B3A239/2020\|EPI_ISL_658817\|20 | ···················· | ···················· | ···················· | ···················· | ···················· | ···················· |
| S | 5341 hCoV-19/England/MILK-B39434/2020\|EPI_ISL_658818\|20 | ···················· | ···················· | ···················· | ···················· | ···················· | ···················· |
| S | 5342 hCoV-19/England/MILK-B393A0/2020\|EPI_ISL_658819\|20 | ···················· | ···················· | ···················· | ···················· | ···················· | ···················· |
| S | 5343 hCoV-19/England/MILK-B38620/2020\|EPI_ISL_658820\|20 | ···················· | ···················· | ···················· | ···················· | ···················· | ···················· |
| S | 5344 hCoV-19/England/MILK-B39540/2020\|EPI_ISL_658821\|20 | ···················· | ···················· | ···················· | ···················· | ···················· | ···················· |
| S | 5345 hCoV-19/England/MILK-B3864E/2020\|EPI_ISL_658822\|20 | ···················· | ···················· | ···················· | ···················· | ···················· | ···················· |
| S | 5346 hCoV-19/England/MILK-B38453/2020\|EPI_ISL_658823\|20 | ···················· | ···················· | ···················· | ···················· | ···················· | ···················· |
| S | 5347 hCoV-19/England/MILK-B38A60/2020\|EPI_ISL_658825\|20 | ···················· | ···················· | ···················· | ···················· | ···················· | ···················· |
| S | 5348 hCoV-19/England/MILK-B394DA/2020\|EPI_ISL_658827\|20 | ···················· | ···················· | ···················· | ···················· | ···················· | ···················· |
| S | 5349 hCoV-19/England/MILK-B38769/2020\|EPI_ISL_658828\|20 | ···················· | ···················· | ···················· | ···················· | ···················· | ···················· |
| S | 5350 hCoV-19/England/CAMC-B37D77/2020\|EPI_ISL_658829\|20 | ···················· | ···················· | ···················· | ···················· | ···················· | ···················· |
| S | 5351 hCoV-19/England/MILK-B38D3A/2020\|EPI_ISL_658830\|20 | ···················· | ···················· | ···················· | ···················· | ···················· | ···················· |
| S | 5352 hCoV-19/England/MILK-B384DB/2020\|EPI_ISL_658831\|20 | ···················· | ···················· | ···················· | ···················· | ···················· | ···················· |
| S | 5353 hCoV-19/England/MILK-B394CB/2020\|EPI_ISL_658832\|20 | ···················· | ···················· | ···················· | ···················· | ···················· | ···················· |
| S | 5354 hCoV-19/England/MILK-B3A293/2020\|EPI_ISL_658834\|20 | ···················· | ···················· | ···················· | ···················· | ···················· | ···················· |
| S | 5355 hCoV-19/England/MILK-B38E91/2020\|EPI_ISL_658835\|20 | ···················· | ···················· | ···················· | ···················· | ···················· | ···················· |
| S | 5356 hCoV-19/England/MILK-B38DD0/2020\|EPI_ISL_658860\|20 | ···················· | ···················· | ···················· | ···················· | ···················· | ···················· |
| S | 5357 hCoV-19/England/CAMC-B380AA/2020\|EPI_ISL_658866\|20 | ···················· | ···················· | ···················· | ···················· | ···················· | ···················· |
| S | 5358 hCoV-19/Mexico/AGS-InDRE-85/2020\|EPI_ISL_658880\|20 | ···················· | ·A·················· | ···················· | ···················· | ···················· | ···················· |
| S | 5359 hCoV-19/England/CAMC-B380E6/2020\|EPI_ISL_658881\|20 | ···················· | ···················· | ···················· | ···················· | ···················· | ···················· |
| S | 5360 hCoV-19/England/MILK-B387C3/2020\|EPI_ISL_658882\|20 | ···················· | ···················· | ···················· | ···················· | ···················· | ···················· |

| | | | | | | | |
|---|---|---|---|---|---|---|---|
| S | 5361 hCoV-19/Mexico/AGS-InDRE-86/2020\|EPI_ISL_658883\|20 | ···················· | ···················· | ···················· | ···················· | ···················· | ···················· |
| S | 5362 hCoV-19/England/MILK-B384CC/2020\|EPI_ISL_658884\|20 | ···················· | ···················· | ···················· | ···················· | ···················· | ···················· |
| S | 5363 hCoV-19/England/CAMC-B38189/2020\|EPI_ISL_658885\|20 | ···················· | ·A·················· | ···················· | ···················· | ···················· | ··A················· |
| S | 5364 hCoV-19/Mexico/QRO-InDRE-87/2020\|EPI_ISL_658886\|20 | ···················· | ···················· | ···················· | ···················· | ···················· | ···················· |
| S | 5365 hCoV-19/England/MILK-B38699/2020\|EPI_ISL_658887\|20 | ···················· | ···················· | ···················· | ···················· | ···················· | ···················· |
| S | 5366 hCoV-19/Mexico/QRO-InDRE-88/2020\|EPI_ISL_658888\|20 | ···················· | ·A·················· | ···················· | ···················· | ···················· | ···················· |
| S | 5367 hCoV-19/England/MILK-B38347/2020\|EPI_ISL_658889\|20 | ···················· | ···················· | ···················· | ···················· | ···················· | ···················· |
| S | 5368 hCoV-19/England/MILK-B38DC1/2020\|EPI_ISL_658890\|20 | ···················· | ···················· | ···················· | ···················· | ···················· | ···················· |
| S | 5369 hCoV-19/Mexico/AGS-InDRE-89/2020\|EPI_ISL_658891\|20 | ···················· | ···················· | ···················· | ···················· | ···················· | ···················· |
| S | 5370 hCoV-19/England/MILK-B39249/2020\|EPI_ISL_658892\|20 | ···················· | ···················· | ···················· | ···················· | ···················· | ···················· |
| S | 5371 hCoV-19/Mexico/QRO-InDRE-90/2020\|EPI_ISL_658893\|20 | ···················· | ···················· | ···················· | ···················· | ···················· | ···················· |
| S | 5372 hCoV-19/England/MILK-B38BF4/2020\|EPI_ISL_658894\|20 | ···················· | ···················· | ···················· | ···················· | ···················· | ···················· |
| S | 5373 hCoV-19/England/MILK-B394BC/2020\|EPI_ISL_658895\|20 | ···················· | ···················· | ···················· | ···················· | ···················· | ···················· |
| S | 5374 hCoV-19/Mexico/GRO-InDRE-91/2020\|EPI_ISL_658896\|20 | ···················· | ···················· | ···················· | ···················· | ···················· | ···················· |
| S | 5375 hCoV-19/England/MILK-B394AD/2020\|EPI_ISL_658897\|20 | ···················· | ···················· | ···················· | ···················· | ···················· | ···················· |
| S | 5376 hCoV-19/England/MILK-B393BF/2020\|EPI_ISL_658898\|20 | ···················· | ···················· | ···················· | ···················· | ···················· | ···················· |
| S | 5377 hCoV-19/USA/MI-UM-10036739832/2020\|EPI_ISL_650001\| | ···················· | ···················· | ···················· | ···················· | ···················· | ···················· |
| S | 5382 hCoV-19/USA/MI-UM-10036620779/2020\|EPI_ISL_650019\| | ···················· | ···················· | ···················· | ···················· | ···················· | ···················· |
| S | 5385 hCoV-19/USA/MI-UM-10036625640/2020\|EPI_ISL_650023\| | ···················· | ···················· | ···················· | ···················· | ···················· | ···················· |
| S | 5387 hCoV-19/USA/MI-UM-10036625470/2020\|EPI_ISL_650030\| | ···················· | ·A·················· | ···················· | ···················· | ···················· | ···················· |
| S | 5388 hCoV-19/USA/MI-UM-10036647686/2020\|EPI_ISL_650031\| | ···················· | ···················· | ···················· | ···················· | ···················· | ···················· |
| S | 5389 hCoV-19/USA/MI-UM-10036625850/2020\|EPI_ISL_650033\| | ···················· | ···················· | ···················· | ···················· | ···················· | ···················· |
| S | 5392 hCoV-19/Wales/PHWC-48A7E7/2020\|EPI_ISL_652600\|2020 | ···················· | ···················· | ···················· | ···················· | ···················· | ···················· |
| S | 5393 hCoV-19/Wales/PHWC-48A7F6/2020\|EPI_ISL_652601\|2020 | ···················· | ···················· | ···················· | ···················· | ···················· | ···················· |
| S | 5394 hCoV-19/Wales/PHWC-48A811/2020\|EPI_ISL_652602\|2020 | ···················· | ···················· | ···················· | ···················· | ···················· | ···················· |
| S | 5395 hCoV-19/Wales/PHWC-48A820/2020\|EPI_ISL_652603\|2020 | ···················· | ···················· | ···················· | ···················· | ···················· | ···················· |
| S | 5396 hCoV-19/Wales/PHWC-48A84E/2020\|EPI_ISL_652604\|2020 | ···················· | ···················· | ···················· | ···················· | ···················· | ···················· |
| S | 5397 hCoV-19/Wales/PHWC-48A85D/2020\|EPI_ISL_652605\|2020 | ···················· | ···················· | ···················· | ···················· | ···················· | ···················· |
| S | 5398 hCoV-19/Wales/PHWC-48A86C/2020\|EPI_ISL_652606\|2020 | ···················· | ···················· | ···················· | ···················· | ···················· | ···················· |
| S | 5399 hCoV-19/Wales/PHWC-48A87B/2020\|EPI_ISL_652607\|2020 | ···················· | ···················· | ···················· | ···················· | ···················· | ···················· |
| S | 5400 hCoV-19/Wales/PHWC-48A88A/2020\|EPI_ISL_652608\|2020 | ···················· | ···················· | ···················· | ···················· | ···················· | ···················· |
| S | 5401 hCoV-19/Wales/PHWC-48A8A8/2020\|EPI_ISL_652609\|2020 | ···················· | ···················· | ···················· | ···················· | ···················· | ···················· |
| S | 5402 hCoV-19/Scotland/QEUH-A9D11A/2020\|EPI_ISL_652654\|2 | ···················· | ···················· | ···················· | ···················· | ···················· | ···················· |
| S | 5403 hCoV-19/England/QEUH-A9D129/2020\|EPI_ISL_652655\|20 | ···················· | ···················· | ···················· | ···················· | ···················· | ···················· |
| S | 5404 hCoV-19/Scotland/QEUH-A9D138/2020\|EPI_ISL_652656\|2 | ···················· | ···················· | ···················· | ···················· | ···················· | ···················· |
| S | 5405 hCoV-19/Scotland/QEUH-A9D147/2020\|EPI_ISL_652657\|2 | ···················· | ···················· | ···················· | ···················· | ···················· | ···················· |
| S | 5406 hCoV-19/Scotland/QEUH-A9D174/2020\|EPI_ISL_652660\|2 | ···················· | ···················· | ···················· | ···················· | ···················· | ···················· |
| S | 5407 hCoV-19/England/QEUH-A9D183/2020\|EPI_ISL_652661\|20 | ···················· | ···················· | ···················· | ···················· | ···················· | ···················· |
| S | 5408 hCoV-19/England/QEUH-A9D97C/2020\|EPI_ISL_652662\|20 | ···················· | ···················· | ···················· | ···················· | ···T················ | ···················· |
| S | 5409 hCoV-19/England/QEUH-A9D98B/2020\|EPI_ISL_652663\|20 | ···················· | ···················· | ···················· | ···················· | ···················· | ···················· |
| S | 5410 hCoV-19/England/QEUH-A9D99A/2020\|EPI_ISL_652664\|20 | ···················· | ···················· | ···················· | ···················· | ···················· | ··A················· |
| S | 5411 hCoV-19/England/QEUH-A9F1DC/2020\|EPI_ISL_652676\|20 | ···················· | ···················· | ···················· | ···················· | ···················· | ···················· |
| S | 5412 hCoV-19/England/QEUH-A9F1FA/2020\|EPI_ISL_652677\|20 | ···················· | ···················· | ···················· | ···················· | ···················· | ···················· |
| S | 5413 hCoV-19/England/QEUH-A9F215/2020\|EPI_ISL_652678\|20 | ···················· | ·A·················· | ···················· | ···················· | ···················· | ··A················· |
| S | 5414 hCoV-19/England/QEUH-A9F233/2020\|EPI_ISL_652679\|20 | ···················· | ···················· | ···················· | ···················· | ···················· | ···················· |
| S | 5415 hCoV-19/England/QEUH-A9F260/2020\|EPI_ISL_652680\|20 | ···················· | ···················· | ···················· | ···················· | ···················· | ···················· |
| S | 5416 hCoV-19/England/QEUH-A9F822/2020\|EPI_ISL_652681\|20 | ···················· | ···················· | ···················· | ···················· | ···················· | ···················· |
| S | 5417 hCoV-19/England/QEUH-A9F840/2020\|EPI_ISL_652682\|20 | ···················· | ···················· | ···················· | ···················· | ···················· | ···················· |
| S | 5418 hCoV-19/England/QEUH-A9F8AA/2020\|EPI_ISL_652683\|20 | ···················· | ···················· | ···················· | ···················· | ···················· | ···················· |
| S | 5419 hCoV-19/England/QEUH-A9F8B9/2020\|EPI_ISL_652684\|20 | ···················· | ···················· | ···················· | ···················· | ···················· | ···················· |
| S | 5420 hCoV-19/England/QEUH-A9F8D7/2020\|EPI_ISL_652685\|20 | ···················· | ···················· | ···················· | ···················· | ···················· | ···················· |
| S | 5421 hCoV-19/England/QEUH-A9F8F5/2020\|EPI_ISL_652686\|20 | ···················· | ···················· | ···················· | ···················· | ···················· | ···················· |
| S | 5422 hCoV-19/England/QEUH-A9F910/2020\|EPI_ISL_652687\|20 | ···················· | ···················· | ···················· | ···················· | ···················· | ···················· |
| S | 5423 hCoV-19/England/QEUH-A9F94D/2020\|EPI_ISL_652688\|20 | ···················· | ···················· | ···················· | ···················· | ···················· | ···················· |
| S | 5424 hCoV-19/Wales/QEUH-A9F9A7/2020\|EPI_ISL_652689\|2020 | ···················· | ···················· | ···················· | ···················· | ···················· | ···················· |
| S | 5425 hCoV-19/England/QEUH-A9F9B6/2020\|EPI_ISL_652690\|20 | ···················· | ·A·················· | ···················· | ···················· | ···················· | ··A················· |
| S | 5426 hCoV-19/England/QEUH-A9FCDB/2020\|EPI_ISL_652691\|20 | ···················· | ·A·················· | ···················· | ···················· | ···················· | ···················· |
| S | 5427 hCoV-19/England/QEUH-A9FCEA/2020\|EPI_ISL_652692\|20 | ···················· | ···················· | ···················· | ···················· | ···················· | ···················· |
| S | 5428 hCoV-19/England/QEUH-A9FD05/2020\|EPI_ISL_652693\|20 | ···················· | ···················· | ···················· | ···················· | ···················· | ···················· |
| S | 5429 hCoV-19/England/QEUH-A9FD23/2020\|EPI_ISL_652694\|20 | ···················· | ···················· | ···················· | ···················· | ···················· | ···················· |
| S | 5430 hCoV-19/England/QEUH-A9FD32/2020\|EPI_ISL_652695\|20 | ···················· | ·A·················· | ···················· | ···················· | ···················· | ···················· |
| S | 5431 hCoV-19/England/QEUH-A9FD41/2020\|EPI_ISL_652696\|20 | ···················· | ···················· | ···················· | ···················· | ···················· | ···················· |
| S | 5432 hCoV-19/England/QEUH-A9FD6F/2020\|EPI_ISL_652697\|20 | ···················· | ···················· | ···················· | ···················· | ···················· | ···················· |
| S | 5433 hCoV-19/India/HR-NIBG-TF84/2020\|EPI_ISL_653900\|202 | ···················· | ···················· | ···················· | ···················· | ···················· | ···················· |
| S | 5434 hCoV-19/India/HR-NIBG-TF85/2020\|EPI_ISL_653901\|202 | ···················· | ···················· | ···················· | ···················· | ···················· | ···················· |
| S | 5435 hCoV-19/India/HR-NIBG-TF86/2020\|EPI_ISL_653902\|202 | ···················· | ···················· | ···················· | ···················· | ···················· | ···················· |
| S | 5436 hCoV-19/India/HR-NIBG-TF88/2020\|EPI_ISL_653904\|202 | ···················· | ·A·················· | ···················· | ···················· | ···················· | ··A················· |
| S | 5437 hCoV-19/India/HR-NIBG-TF89/2020\|EPI_ISL_653905\|202 | ···················· | ·A·················· | ···················· | ···················· | ···················· | ··A················· |
| S | 5438 hCoV-19/India/HR-NIBG-TF91/2020\|EPI_ISL_653906\|202 | ···················· | ···················· | ···················· | ···················· | ···················· | ···················· |
| S | 5439 hCoV-19/India/HR-NIBG-TF93/2020\|EPI_ISL_653908\|202 | ···················· | ···················· | ···················· | ···················· | ···················· | ···················· |
| S | 5440 hCoV-19/India/HR-NIBG-TF94/2020\|EPI_ISL_653909\|202 | ···················· | ···················· | ···················· | ···················· | ···················· | ···················· |
| S | 5441 hCoV-19/India/HR-NIBG-TF95/2020\|EPI_ISL_653910\|202 | ···················· | ·A·················· | ···················· | ···················· | ···················· | ··A················· |
| S | 5442 hCoV-19/India/HR-NIBG-TF96/2020\|EPI_ISL_653911\|202 | ···················· | ·A·················· | ···················· | ···················· | ···················· | ··A················· |
| S | 5443 hCoV-19/India/HR-NIBG-TF97/2020\|EPI_ISL_653912\|202 | ···················· | ···················· | ···················· | ···················· | ···················· | ···················· |
| S | 5444 hCoV-19/India/HR-NIBG-TF98/2020\|EPI_ISL_653913\|202 | ···················· | ·A·················· | ···················· | ···················· | ···················· | ··A················· |
| S | 5445 hCoV-19/India/HR-NIBG-TF99/2020\|EPI_ISL_653914\|202 | ···················· | ·A·················· | ···················· | ···················· | ···················· | ··A················· |
| S | 5446 hCoV-19/Austria/Graz-MUG20/2020\|EPI_ISL_653916\|202 | ···················· | ···················· | ···················· | ···················· | ···················· | ···················· |
| S | 5447 hCoV-19/Russia/MOS-CRIE7162018/2020\|EPI_ISL_653922 | ···················· | ···················· | ···················· | ···················· | ···················· | ···················· |
| S | 5448 hCoV-19/Russia/MOS-CRIE7160676/2020\|EPI_ISL_653923 | ···················· | ···················· | ···················· | ···················· | ···················· | ···················· |
| S | 5449 hCoV-19/Russia/MOS-CRIE7150916/2020\|EPI_ISL_653924 | ···················· | ···················· | ···················· | ···················· | ···················· | ···················· |

| | | | | | | | |
|---|---|---|---|---|---|---|---|
| S | 5450 hCoV-19/Russia/MOS-CRIE7150909/2020\|EPI_ISL_653925 | ···················· | ···················· | ····················· | ···················· | ···················· | ···················· |
| S | 5451 hCoV-19/Russia/MOS-CRIE7169887/2020\|EPI_ISL_653926 | ···················· | ···················· | ····················· | ···················· | ···················· | ···················· |
| S | 5452 hCoV-19/Russia/MOS-CRIE7129338/2020\|EPI_ISL_653927 | ···················· | ···················· | ····················· | ···················· | ···················· | ···················· |
| S | 5453 hCoV-19/Russia/MOS-CRIE7122594/2020\|EPI_ISL_653928 | ···················· | ···················· | ····················· | ···················· | ···················· | ···················· |
| S | 5454 hCoV-19/Russia/MOS-CRIE7150932/2020\|EPI_ISL_653929 | ···················· | ···················· | ····················· | ···················· | ···················· | ···················· |
| S | 5455 hCoV-19/Russia/MOS-CRIE7165818/2020\|EPI_ISL_653930 | ···················· | ···················· | ····················· | ···················· | ···················· | ···················· |
| S | 5456 hCoV-19/Russia/MOS-CRIE7157442/2020\|EPI_ISL_653931 | ···················· | ···················· | ····················· | ···················· | ···················· | ···················· |
| S | 5457 hCoV-19/Russia/MOS-CRIE7141819/2020\|EPI_ISL_653932 | ···················· | ···················· | ····················· | ···················· | ···················· | ···················· |
| S | 5458 hCoV-19/England/204462384/2020\|EPI_ISL_653985\|2020 | ···················· | ···················· | ····················· | ···················· | ···················· | ···················· |
| S | 5459 hCoV-19/England/204462584/2020\|EPI_ISL_653986\|2020 | ···················· | ···················· | ····················· | ···················· | ···················· | ···················· |
| S | 5460 hCoV-19/England/204462588/2020\|EPI_ISL_653987\|2020 | ···················· | ···················· | ····················· | ···················· | ···················· | ···················· |
| S | 5461 hCoV-19/England/204462610/2020\|EPI_ISL_653988\|2020 | ···················· | ···················· | ····················· | ···················· | ···················· | ···················· |
| S | 5462 hCoV-19/England/204481768/2020\|EPI_ISL_653990\|2020 | ···················· | ···················· | ····················· | ···················· | ···················· | ···················· |
| S | 5463 hCoV-19/England/204481773/2020\|EPI_ISL_653991\|2020 | ···················· | ···················· | ····················· | ···················· | ···················· | ···················· |
| S | 5464 hCoV-19/England/204481774/2020\|EPI_ISL_653992\|2020 | ···················· | ···················· | ····················· | ···················· | ···················· | ···················· |
| S | 5465 hCoV-19/England/204481780/2020\|EPI_ISL_653993\|2020 | ···················· | ···················· | ····················· | ···················· | ···················· | ···················· |
| S | 5466 hCoV-19/England/204481791/2020\|EPI_ISL_653994\|2020 | ···················· | ···················· | ····················· | ···················· | ···················· | ···················· |
| S | 5467 hCoV-19/England/204481795/2020\|EPI_ISL_653995\|2020 | ···················· | ···················· | ····················· | ···················· | ···················· | ···················· |
| S | 5468 hCoV-19/England/ALDP-B46FAC/2020\|EPI_ISL_656140\|20 | ···················· | ···················· | ····················· | ···················· | ···················· | ···················· |
| S | 5469 hCoV-19/England/ALDP-B469DB/2020\|EPI_ISL_656141\|20 | ···················· | ···················· | ····················· | ···················· | ···················· | ···················· |
| S | 5470 hCoV-19/England/ALDP-B46A41/2020\|EPI_ISL_656143\|20 | ···················· | ···················· | ····················· | ···················· | ···················· | ···················· |
| S | 5471 hCoV-19/England/ALDP-B46698/2020\|EPI_ISL_656144\|20 | ···················· | ···················· | ····················· | ···················· | ···················· | ···················· |
| S | 5472 hCoV-19/England/ALDP-B45A7F/2020\|EPI_ISL_656145\|20 | ···················· | ···················· | ····················· | ···················· | ···················· | ···················· |
| S | 5473 hCoV-19/England/ALDP-B46C0F/2020\|EPI_ISL_656146\|20 | ···················· | ·A·················· | ····················· | ···················· | ···················· | ···················· |
| S | 5474 hCoV-19/England/ALDP-B45945/2020\|EPI_ISL_656147\|20 | ···················· | ···················· | ····················· | ···················· | ···················· | ···················· |
| S | 5475 hCoV-19/England/ALDP-B45FBC/2020\|EPI_ISL_656148\|20 | ···················· | ···················· | ····················· | ···················· | ···················· | ···················· |
| S | 5476 hCoV-19/England/ALDP-B45DFE/2020\|EPI_ISL_656149\|20 | ···················· | ···················· | ····················· | ···················· | ···················· | ···················· |
| S | 5477 hCoV-19/England/ALDP-B46D2A/2020\|EPI_ISL_656150\|20 | ···················· | ···················· | ····················· | ···················· | ···················· | ···················· |
| S | 5478 hCoV-19/England/ALDP-B4666B/2020\|EPI_ISL_656151\|20 | ···················· | ···················· | ····················· | ···················· | ···················· | ···················· |
| S | 5479 hCoV-19/England/ALDP-B469EA/2020\|EPI_ISL_656152\|20 | ···················· | ···················· | ····················· | ···················· | ···················· | ···················· |
| S | 5480 hCoV-19/England/ALDP-B46CB4/2020\|EPI_ISL_656153\|20 | ···················· | ···················· | ····················· | ···················· | ···················· | ···················· |
| S | 5481 hCoV-19/England/ALDP-B472EE/2020\|EPI_ISL_656154\|20 | ···················· | ·A·················· | ····················· | ···················· | ···················· | ···················· |
| S | 5482 hCoV-19/England/ALDP-B46AF6/2020\|EPI_ISL_656155\|20 | ···················· | ···················· | ····················· | ···················· | ···················· | ···················· |
| S | 5483 hCoV-19/England/ALDP-B467B3/2020\|EPI_ISL_656156\|20 | ···················· | ···················· | ····················· | ···················· | ···················· | ···················· |
| S | 5484 hCoV-19/England/ALDP-B45990/2020\|EPI_ISL_656157\|20 | ···················· | ···················· | ····················· | ···················· | ···················· | ···················· |
| S | 5485 hCoV-19/England/ALDP-B466B6/2020\|EPI_ISL_656158\|20 | ···················· | ···················· | ····················· | ···················· | ···················· | ···················· |
| S | 5486 hCoV-19/England/ALDP-B472C0/2020\|EPI_ISL_656159\|20 | ···················· | ·A·················· | ····················· | ···················· | ···················· | ···················· |
| S | 5487 hCoV-19/England/ALDP-B46F9D/2020\|EPI_ISL_656160\|20 | ···················· | ···················· | ····················· | ···················· | ···················· | ···················· |
| S | 5488 hCoV-19/England/ALDP-B464AD/2020\|EPI_ISL_656162\|20 | ···················· | ···················· | ····················· | ···················· | ···················· | ···················· |
| S | 5489 hCoV-19/England/ALDP-B46C78/2020\|EPI_ISL_656165\|20 | ···················· | ···················· | ····················· | ···················· | ···················· | ···················· |
| S | 5490 hCoV-19/England/ALDP-B45E55/2020\|EPI_ISL_656166\|20 | ···················· | ···················· | ····················· | ···················· | ···················· | ···················· |
| S | 5491 hCoV-19/England/ALDP-B46012/2020\|EPI_ISL_656167\|20 | ···················· | ···················· | ····················· | ···················· | ···················· | ···················· |
| S | 5492 hCoV-19/England/ALDP-B45B8B/2020\|EPI_ISL_656168\|20 | ···················· | ···················· | ····················· | ···················· | ···················· | ···················· |
| S | 5493 hCoV-19/England/ALDP-B4608B/2020\|EPI_ISL_656169\|20 | ···················· | ···················· | ····················· | ···················· | ···················· | ···················· |
| S | 5494 hCoV-19/England/ALDP-B45F16/2020\|EPI_ISL_656170\|20 | ···················· | ···················· | ····················· | ···················· | ···················· | ···················· |
| S | 5495 hCoV-19/England/ALDP-B460A9/2020\|EPI_ISL_656171\|20 | ···················· | ···················· | ····················· | ···················· | ···················· | ···················· |
| S | 5496 hCoV-19/England/ALDP-B467FF/2020\|EPI_ISL_656172\|20 | ···················· | ···················· | ····················· | ···················· | ···················· | ···················· |
| S | 5497 hCoV-19/England/ALDP-B45A15/2020\|EPI_ISL_656175\|20 | ···················· | ···················· | ····················· | ···················· | ···················· | ···················· |
| S | 5498 hCoV-19/England/ALDP-B467D1/2020\|EPI_ISL_656176\|20 | ···················· | ···················· | ····················· | ···················· | ···················· | ···················· |
| S | 5499 hCoV-19/England/ALDP-B466A7/2020\|EPI_ISL_656178\|20 | ···················· | ···················· | ····················· | ···················· | ···················· | ···················· |
| S | 5500 hCoV-19/England/ALDP-B46CF0/2020\|EPI_ISL_656179\|20 | ···················· | ···················· | ····················· | ···················· | ···················· | ···················· |
| S | 5501 hCoV-19/England/ALDP-B45C6A/2020\|EPI_ISL_656181\|20 | ···················· | ···················· | ····················· | ···················· | ···················· | ···················· |
| S | 5502 hCoV-19/England/ALDP-B472B1/2020\|EPI_ISL_656182\|20 | ···················· | ·A·················· | ····················· | ···················· | ···················· | ···················· |
| S | 5503 hCoV-19/England/ALDP-B46A8D/2020\|EPI_ISL_656183\|20 | ···················· | ···················· | ····················· | ···················· | ···················· | ···················· |
| S | 5504 hCoV-19/England/ALDP-B45A42/2020\|EPI_ISL_656184\|20 | ···················· | ···················· | ····················· | ···················· | ···················· | ···················· |
| S | 5505 hCoV-19/England/ALDP-B46E54/2020\|EPI_ISL_656185\|20 | ···················· | ···················· | ····················· | ···················· | ···················· | ···················· |
| S | 5506 hCoV-19/England/ALDP-B45E82/2020\|EPI_ISL_656186\|20 | ···················· | ···················· | ····················· | ···················· | ···················· | ···················· |
| S | 5507 hCoV-19/England/ALDP-B45E28/2020\|EPI_ISL_656187\|20 | ···················· | ···················· | ····················· | ···················· | ···················· | ···················· |
| S | 5508 hCoV-19/England/ALDP-B462D0/2020\|EPI_ISL_656188\|20 | ···················· | ···················· | ····················· | ···················· | ···················· | ···················· |
| S | 5509 hCoV-19/England/ALDP-B46258/2020\|EPI_ISL_656189\|20 | ···················· | ···················· | ····················· | ···················· | ···················· | ···················· |
| S | 5510 hCoV-19/England/ALDP-B46FCA/2020\|EPI_ISL_656191\|20 | ···················· | ···················· | ····················· | ···················· | ···················· | ···················· |
| S | 5511 hCoV-19/England/ALDP-B45A51/2020\|EPI_ISL_656193\|20 | ···················· | ···················· | ····················· | ···················· | ···················· | ···················· |
| S | 5512 hCoV-19/England/ALDP-B4623A/2020\|EPI_ISL_656194\|20 | ···················· | ···················· | ····················· | ···················· | ···················· | ···················· |
| S | 5513 hCoV-19/England/ALDP-B46AC9/2020\|EPI_ISL_656195\|20 | ···················· | ···················· | ····················· | ···················· | ···················· | ···················· |
| S | 5514 hCoV-19/England/ALDP-B47327/2020\|EPI_ISL_656196\|20 | ···················· | ···················· | ····················· | ···················· | ···················· | ···················· |
| S | 5515 hCoV-19/England/ALDP-B465C8/2020\|EPI_ISL_656197\|20 | ···················· | ···················· | ····················· | ···················· | ···················· | ···················· |
| S | 5516 hCoV-19/England/ALDP-B46A6F/2020\|EPI_ISL_656198\|20 | ···················· | ···················· | ····················· | ···················· | ···················· | ···················· |
| S | 5517 hCoV-19/England/ALDP-B45A24/2020\|EPI_ISL_656199\|20 | ···················· | ···················· | ····················· | ···················· | ···················· | ···················· |
| S | 5518 hCoV-19/England/ALDP-B46249/2020\|EPI_ISL_656200\|20 | ···················· | ···················· | ····················· | ···················· | ···················· | ···················· |
| S | 5519 hCoV-19/England/ALDP-B46D1B/2020\|EPI_ISL_656201\|20 | ···················· | ···················· | ····················· | ···················· | ···················· | ···················· |
| S | 5520 hCoV-19/England/ALDP-B45A33/2020\|EPI_ISL_656202\|20 | ···················· | ···················· | ····················· | ···················· | ···················· | ···················· |
| S | 5521 hCoV-19/England/ALDP-B46F60/2020\|EPI_ISL_656204\|20 | ···················· | ···················· | ····················· | ···················· | ···················· | ···················· |
| S | 5522 hCoV-19/England/ALDP-B46179/2020\|EPI_ISL_656205\|20 | ···················· | ···················· | ····················· | ···················· | ···················· | ···················· |
| S | 5523 hCoV-19/England/ALDP-B460B8/2020\|EPI_ISL_656206\|20 | ···················· | ···················· | ····················· | ···················· | ···················· | ···················· |
| S | 5524 hCoV-19/England/ALDP-B45CF1/2020\|EPI_ISL_656207\|20 | ···················· | ···················· | ····················· | ···················· | ···················· | ···················· |
| S | 5525 hCoV-19/England/ALDP-B47309/2020\|EPI_ISL_656208\|20 | ···················· | ···················· | ····················· | ···················· | ···················· | ···················· |
| S | 5526 hCoV-19/England/ALDP-B45C2E/2020\|EPI_ISL_656209\|20 | ···················· | ···················· | ····················· | ···················· | ···················· | ···················· |
| S | 5527 hCoV-19/England/ALDP-B463BF/2020\|EPI_ISL_656210\|20 | ···················· | ·A·················· | ····················· | ···················· | ···················· | ···················· |
| S | 5528 hCoV-19/England/ALDP-B47345/2020\|EPI_ISL_656211\|20 | ···················· | ···················· | ····················· | ···················· | ···················· | ···················· |
| S | 5529 hCoV-19/England/ALDP-B45C97/2020\|EPI_ISL_656212\|20 | ···················· | ···················· | ····················· | ···················· | ···················· | ···················· |

| | | | | | | | |
|---|---|---|---|---|---|---|---|
| S | 5530 hCoV-19/England/ALDP-B45A8E/2020\|EPI_ISL_656213\|20 | ···················· | ···················· | ···················· | ···················· | ···················· | ···················· |
| S | 5531 hCoV-19/England/ALDP-B4670E/2020\|EPI_ISL_656214\|20 | ···················· | ···················· | ···················· | ···················· | ···················· | ···················· |
| S | 5532 hCoV-19/England/ALDP-B46E09/2020\|EPI_ISL_656215\|20 | ···················· | ···················· | ···················· | ···················· | ···················· | ···················· |
| S | 5533 hCoV-19/England/ALDP-B46319/2020\|EPI_ISL_656216\|20 | ···················· | ···················· | ···················· | ···················· | ···················· | ···················· |
| S | 5534 hCoV-19/England/ALDP-B45F52/2020\|EPI_ISL_656217\|20 | ···················· | ···················· | ···················· | ···················· | ···················· | ···················· |
| S | 5535 hCoV-19/England/CAMC-B52C79/2020\|EPI_ISL_657470\|20 | ···················· | ···················· | ···················· | ···················· | ···················· | ···················· |
| S | 5536 hCoV-19/England/CAMC-B530A9/2020\|EPI_ISL_657471\|20 | ···················· | ···················· | ···················· | ···················· | ···················· | ···················· |
| S | 5537 hCoV-19/England/MILK-B5565A/2020\|EPI_ISL_657472\|20 | ···················· | ···················· | ···················· | ···················· | ···················· | ···················· |
| S | 5538 hCoV-19/England/MILK-B561C1/2020\|EPI_ISL_657473\|20 | ···················· | ···················· | ···················· | ···················· | ···················· | ···················· |
| S | 5539 hCoV-19/England/CAMC-B52D58/2020\|EPI_ISL_657474\|20 | ···················· | ···················· | ···················· | ···················· | ···················· | ···················· |
| S | 5540 hCoV-19/England/CAMC-B52DA3/2020\|EPI_ISL_657475\|20 | ···················· | ···················· | ···················· | ···················· | ···················· | ···················· |
| S | 5541 hCoV-19/England/CAMC-B52C97/2020\|EPI_ISL_657477\|20 | ···················· | ···················· | ···················· | ···················· | ···················· | ···················· |
| S | 5542 hCoV-19/England/MILK-B567DE/2020\|EPI_ISL_657479\|20 | ···················· | ···················· | ···················· | ···················· | ···················· | ···················· |
| S | 5543 hCoV-19/England/CAMC-B52927/2020\|EPI_ISL_657481\|20 | ···················· | ···················· | ···················· | ···················· | ···················· | ···················· |
| S | 5544 hCoV-19/England/CAMC-B53346/2020\|EPI_ISL_657482\|20 | ···················· | ···················· | ···················· | ···················· | ···················· | ···················· |
| S | 5545 hCoV-19/England/MILK-B563F8/2020\|EPI_ISL_657483\|20 | ···················· | ···················· | ···················· | ···················· | ···················· | ···A················ |
| S | 5546 hCoV-19/England/MILK-B566A4/2020\|EPI_ISL_657484\|20 | ···················· | ···················· | ···················· | ···················· | ···················· | ···················· |
| S | 5547 hCoV-19/England/MILK-B55C0D/2020\|EPI_ISL_657487\|20 | ···················· | ···················· | ···················· | ···················· | ······T············· | ···················· |
| S | 5548 hCoV-19/England/MILK-B55687/2020\|EPI_ISL_657488\|20 | ···················· | ···················· | ···················· | ···················· | ···················· | ···················· |
| S | 5549 hCoV-19/England/CAMC-B52C4C/2020\|EPI_ISL_657489\|20 | ···················· | ···················· | ···················· | ···················· | ···················· | ···················· |
| S | 5550 hCoV-19/England/MILK-B55B88/2020\|EPI_ISL_657490\|20 | ···················· | ···················· | ···················· | ···················· | ···················· | ···················· |
| S | 5551 hCoV-19/England/MILK-B55739/2020\|EPI_ISL_657491\|20 | ···················· | ···················· | ···················· | ···················· | ···················· | ···················· |
| S | 5552 hCoV-19/England/MILK-B56264/2020\|EPI_ISL_657493\|20 | ···················· | ···················· | ···················· | ···················· | ···················· | ···················· |
| S | 5553 hCoV-19/England/CAMC-B52848/2020\|EPI_ISL_657496\|20 | ···················· | ···················· | ···················· | ···················· | ···················· | ···················· |
| S | 5554 hCoV-19/England/MILK-B55B97/2020\|EPI_ISL_657497\|20 | ···················· | ···················· | ···················· | ···················· | ···················· | ···················· |
| S | 5555 hCoV-19/England/CAMC-B5313D/2020\|EPI_ISL_657498\|20 | ···················· | ···················· | ···················· | ···················· | ···················· | ···················· |
| S | 5556 hCoV-19/England/CAMC-B530E5/2020\|EPI_ISL_657499\|20 | ···················· | ···················· | ···················· | ···················· | ···················· | ···················· |
| S | 5557 hCoV-19/England/MILK-B55B4C/2020\|EPI_ISL_657505\|20 | ···················· | ···················· | ···················· | ···················· | ···················· | ···················· |
| S | 5558 hCoV-19/England/CAMC-B52A06/2020\|EPI_ISL_657506\|20 | ···················· | ·A·················· | ···················· | ···················· | ···················· | ···A················ |
| S | 5559 hCoV-19/England/CAMC-B52CD3/2020\|EPI_ISL_657507\|20 | ···················· | ···················· | ···················· | ···················· | ···················· | ···················· |
| S | 5560 hCoV-19/England/CAMC-B529DC/2020\|EPI_ISL_657508\|20 | ···················· | ···················· | ···················· | ···················· | ···················· | ···················· |
| S | 5561 hCoV-19/England/MILK-B556E1/2020\|EPI_ISL_657509\|20 | ···················· | ···················· | ···················· | ···················· | ···················· | ···················· |
| S | 5562 hCoV-19/England/MILK-B5655C/2020\|EPI_ISL_657510\|20 | ···················· | ···················· | ···················· | ···················· | ···················· | ···················· |
| S | 5563 hCoV-19/England/MILK-B5645F/2020\|EPI_ISL_657512\|20 | ···················· | ···················· | ···················· | ···················· | ···················· | ···················· |
| S | 5564 hCoV-19/England/MILK-B55881/2020\|EPI_ISL_657513\|20 | ···················· | ·A·················· | ···················· | ···················· | ···················· | ···A················ |
| S | 5565 hCoV-19/England/CAMC-B5314C/2020\|EPI_ISL_657514\|20 | ···················· | ···················· | ···················· | ···················· | ···················· | ···················· |
| S | 5566 hCoV-19/England/MILK-B564F5/2020\|EPI_ISL_657515\|20 | ···················· | ···················· | ···················· | ···················· | ···················· | ···················· |
| S | 5567 hCoV-19/England/CAMC-B52A7F/2020\|EPI_ISL_657516\|20 | ···················· | ···················· | ···················· | ···················· | ···················· | ···················· |
| S | 5568 hCoV-19/England/MILK-B564AA/2020\|EPI_ISL_657518\|20 | ···················· | ···················· | ···················· | ···················· | ···················· | ···················· |
| S | 5569 hCoV-19/England/MILK-B562EC/2020\|EPI_ISL_657519\|20 | ···················· | ·A·················· | ···················· | ···················· | ···················· | ···A················ |
| S | 5570 hCoV-19/England/CAMC-B527C3/2020\|EPI_ISL_657520\|20 | ···················· | ···················· | ···················· | ···················· | ···················· | ···················· |
| S | 5571 hCoV-19/England/CAMC-B52DFE/2020\|EPI_ISL_657522\|20 | ···················· | ···················· | ···················· | ···················· | ···················· | ···················· |
| S | 5572 hCoV-19/England/CAMC-B53285/2020\|EPI_ISL_657523\|20 | ···················· | ···················· | ···················· | ···················· | ···················· | ···················· |
| S | 5573 hCoV-19/England/CAMC-B5280C/2020\|EPI_ISL_657524\|20 | ···················· | ···················· | ···················· | ···················· | ···················· | ···················· |
| S | 5574 hCoV-19/England/CAMC-B52A60/2020\|EPI_ISL_657525\|20 | ···················· | ···················· | ···················· | ···················· | ······T············· | ···············A···· |
| S | 5575 hCoV-19/England/MILK-B565A7/2020\|EPI_ISL_657526\|20 | ···················· | ···················· | ···················· | ···················· | ···················· | ···················· |
| S | 5576 hCoV-19/England/MILK-B5599D/2020\|EPI_ISL_657527\|20 | ···················· | ···················· | ···················· | ···················· | ···················· | ···················· |
| S | 5577 hCoV-19/England/CAMC-B53197/2020\|EPI_ISL_657528\|20 | ···················· | ···················· | ···················· | ···················· | ···················· | ···················· |
| S | 5578 hCoV-19/England/CAMC-B52ECE/2020\|EPI_ISL_657529\|20 | ···················· | ···················· | ···················· | ···················· | ···················· | ···················· |
| S | 5579 hCoV-19/England/CAMC-B52E55/2020\|EPI_ISL_657530\|20 | ···················· | ···················· | ···················· | ···················· | ···················· | ···················· |
| S | 5580 hCoV-19/England/MILK-B567ED/2020\|EPI_ISL_657531\|20 | ···················· | ·A·················· | ···················· | ···················· | ···················· | ···················· |
| S | 5581 hCoV-19/England/MILK-B556A5/2020\|EPI_ISL_657532\|20 | ···················· | ···················· | ···················· | ···················· | ···················· | ···················· |
| S | 5582 hCoV-19/England/MILK-B55890/2020\|EPI_ISL_657533\|20 | ···················· | ···················· | ···················· | ···················· | ···················· | ···················· |
| S | 5583 hCoV-19/England/CAMC-B53179/2020\|EPI_ISL_657534\|20 | ···················· | ···················· | ···················· | ···················· | ···················· | ···················· |
| S | 5584 hCoV-19/England/MILK-B5662C/2020\|EPI_ISL_657535\|20 | ···················· | ···················· | ···················· | ···················· | ···················· | ···················· |
| S | 5585 hCoV-19/England/MILK-B5670B/2020\|EPI_ISL_657536\|20 | ···················· | ···················· | ···················· | ···················· | ···················· | ···················· |
| S | 5586 hCoV-19/England/CAMC-B52866/2020\|EPI_ISL_657537\|20 | ···················· | ···················· | ···················· | ···················· | ···················· | ···················· |
| S | 5587 hCoV-19/England/CAMC-B52C2E/2020\|EPI_ISL_657538\|20 | ···················· | ···················· | ···················· | ···················· | ······T············· | ···················· |
| S | 5588 hCoV-19/England/CAMC-B52BE5/2020\|EPI_ISL_657540\|20 | ···················· | ···················· | ···················· | ···················· | ···················· | ···················· |
| S | 5589 hCoV-19/England/CAMC-B52E73/2020\|EPI_ISL_657541\|20 | ···················· | ···················· | ···················· | ···················· | ···················· | ···················· |
| S | 5590 hCoV-19/England/MILK-B556B4/2020\|EPI_ISL_657542\|20 | ···················· | ···················· | ···················· | ···················· | ···················· | ···················· |
| S | 5591 hCoV-19/England/MILK-B563BC/2020\|EPI_ISL_657543\|20 | ···················· | ···················· | ···················· | ···················· | ···················· | ···················· |
| S | 5592 hCoV-19/England/MILK-B55B79/2020\|EPI_ISL_657544\|20 | ···················· | ···················· | ···················· | ···················· | ···················· | ···················· |
| S | 5593 hCoV-19/England/CAMC-B52F16/2020\|EPI_ISL_657545\|20 | ···················· | ···················· | ···················· | ···················· | ···················· | ···················· |
| S | 5594 hCoV-19/England/CAMC-B5306D/2020\|EPI_ISL_657546\|20 | ···················· | ···················· | ···················· | ···················· | ···················· | ···················· |
| S | 5595 hCoV-19/England/MILK-B56589/2020\|EPI_ISL_657547\|20 | ···················· | ···················· | ···················· | ···················· | ···················· | ···················· |
| S | 5596 hCoV-19/England/CAMC-B37CF2/2020\|EPI_ISL_658836\|20 | ···················· | ···················· | ···················· | ···················· | ···················· | ···················· |
| S | 5597 hCoV-19/England/CAMC-B3808C/2020\|EPI_ISL_658837\|20 | ···················· | ·A·················· | ···················· | ···················· | ···················· | ···················· |
| S | 5598 hCoV-19/England/CAMC-B37E0B/2020\|EPI_ISL_658838\|20 | ···················· | ···················· | ···················· | ···················· | ···················· | ···················· |
| S | 5599 hCoV-19/England/MILK-B385AB/2020\|EPI_ISL_658839\|20 | ···················· | ···················· | ···················· | ···················· | ···················· | ···················· |
| S | 5600 hCoV-19/England/MILK-B38E55/2020\|EPI_ISL_658841\|20 | ···················· | ···················· | ···················· | ···················· | ···················· | ···················· |
| S | 5601 hCoV-19/England/MILK-B38444/2020\|EPI_ISL_658842\|20 | ···················· | ···················· | ···················· | ···················· | ···················· | ···················· |
| S | 5602 hCoV-19/England/MILK-B38602/2020\|EPI_ISL_658843\|20 | ···················· | ···················· | ···················· | ···················· | ···················· | ···················· |
| S | 5603 hCoV-19/England/MILK-B39452/2020\|EPI_ISL_658845\|20 | ···················· | ···················· | ···················· | ···················· | ···················· | ···················· |
| S | 5604 hCoV-19/England/CAMC-B37EDE/2020\|EPI_ISL_658846\|20 | ···················· | ···················· | ···················· | ···················· | ···················· | ···················· |
| S | 5605 hCoV-19/England/MILK-B395AA/2020\|EPI_ISL_658847\|20 | ···················· | ···················· | ···················· | ···················· | ···················· | ···················· |
| S | 5606 hCoV-19/England/MILK-B38A7F/2020\|EPI_ISL_658849\|20 | ···················· | ···················· | ···················· | ···················· | ···················· | ···················· |
| S | 5607 hCoV-19/England/MILK-B38462/2020\|EPI_ISL_658850\|20 | ···················· | ···················· | ···················· | ···················· | ···················· | ···················· |
| S | 5608 hCoV-19/England/MILK-B395E6/2020\|EPI_ISL_658851\|20 | ···················· | ···················· | ···················· | ···················· | ···················· | ···················· |
| S | 5609 hCoV-19/England/CAMC-B3813E/2020\|EPI_ISL_658852\|20 | ···················· | ···················· | ···················· | ···················· | ···················· | ···················· |

| | | | | | | | |
|---|---|---|---|---|---|---|---|
| S | 5610 hCoV-19/England/MILK-B38338/2020\|EPI_ISL_658853\|20 | ···················· | ···················· | ····················· | ···················· | ···················· | ···················· |
| S | 5611 hCoV-19/England/MILK-B383ED/2020\|EPI_ISL_658854\|20 | ···················· | ···················· | ····················· | ···················· | ···················· | ···················· |
| S | 5612 hCoV-19/England/MILK-B3959B/2020\|EPI_ISL_658855\|20 | ···················· | ·A·················· | ····················· | ···················· | ···················· | ··A················· |
| S | 5613 hCoV-19/England/MILK-B3865D/2020\|EPI_ISL_658857\|20 | ···················· | ···················· | ····················· | ···················· | ···················· | ···················· |
| S | 5614 hCoV-19/England/MILK-B391B5/2020\|EPI_ISL_658858\|20 | ···················· | ···················· | ····················· | ···················· | ···················· | ···················· |
| S | 5615 hCoV-19/England/MILK-B38C2E/2020\|EPI_ISL_658861\|20 | ···················· | ···················· | ····················· | ···················· | ···················· | ···················· |
| S | 5616 hCoV-19/England/MILK-B38B03/2020\|EPI_ISL_658862\|20 | ···················· | ···················· | ····················· | ···················· | ···················· | ···················· |
| S | 5617 hCoV-19/Mexico/AGS-InDRE-78/2020\|EPI_ISL_658863\|20 | ···················· | ···················· | ····················· | ···················· | ···················· | ···················· |
| S | 5618 hCoV-19/England/CAMC-B381E3/2020\|EPI_ISL_658864\|20 | ···················· | ···················· | ····················· | ···················· | ······T············· | ···················· |
| S | 5619 hCoV-19/Mexico/AGS-InDRE-79/2020\|EPI_ISL_658865\|20 | ···················· | ·A·················· | ····················· | ···················· | ···················· | ···················· |
| S | 5620 hCoV-19/England/MILK-B387E1/2020\|EPI_ISL_658867\|20 | ···················· | ···················· | ····················· | ···················· | ···················· | ···················· |
| S | 5621 hCoV-19/Mexico/AGS-InDRE-80/2020\|EPI_ISL_658868\|20 | ···················· | ···················· | ····················· | ···················· | ···················· | ···················· |
| S | 5622 hCoV-19/England/MILK-B38990/2020\|EPI_ISL_658869\|20 | ···················· | ···················· | ····················· | ···················· | ···················· | ···················· |
| S | 5623 hCoV-19/Mexico/AGS-InDRE-81/2020\|EPI_ISL_658870\|20 | ···················· | ·A·················· | ····················· | ···················· | ···················· | ···················· |
| S | 5624 hCoV-19/England/MILK-B39698/2020\|EPI_ISL_658871\|20 | ···················· | ···················· | ····················· | ···················· | ···················· | ···················· |
| S | 5625 hCoV-19/England/MILK-B39382/2020\|EPI_ISL_658872\|20 | ···················· | ···················· | ····················· | ···················· | ···················· | ···················· |
| S | 5626 hCoV-19/Mexico/AGS-InDRE-82/2020\|EPI_ISL_658873\|20 | ···················· | ···················· | ····················· | ···················· | ···················· | ···················· |
| S | 5627 hCoV-19/England/MILK-B39407/2020\|EPI_ISL_658874\|20 | ···················· | ···················· | ····················· | ···················· | ···················· | ···················· |
| S | 5628 hCoV-19/Mexico/AGS-InDRE-83/2020\|EPI_ISL_658875\|20 | ···················· | ···················· | ····················· | ···················· | ···················· | ···················· |
| S | 5629 hCoV-19/England/MILK-B3A20C/2020\|EPI_ISL_658876\|20 | ···················· | ···················· | ····················· | ···················· | ···················· | ···················· |
| S | 5630 hCoV-19/England/CAMC-B38286/2020\|EPI_ISL_658877\|20 | ···················· | ···················· | ····················· | ···················· | ···················· | ···················· |
| S | 5631 hCoV-19/Mexico/QRO-InDRE-84/2020\|EPI_ISL_658878\|20 | ···················· | ···················· | ····················· | ···················· | ···················· | ···················· |
| S | 5632 hCoV-19/England/MILK-B3956E/2020\|EPI_ISL_658879\|20 | ···················· | ···················· | ····················· | ···················· | ···················· | ···················· |
| S | 5637 hCoV-19/USA/MI-UM-10036587651/2020\|EPI_ISL_650008\| | ···················· | ···················· | ····················· | ···················· | ···················· | ···················· |
| S | 5638 hCoV-19/USA/MI-UM-10036593790/2020\|EPI_ISL_650011\| | ···················· | ···················· | ····················· | ···················· | ···················· | ···················· |
| S | 5640 hCoV-19/Wales/PHWC-48A626/2020\|EPI_ISL_652577\|2020 | ···················· | ···················· | ····················· | ···················· | ···················· | ···················· |
| S | 5641 hCoV-19/Wales/PHWC-48A635/2020\|EPI_ISL_652578\|2020 | ···················· | ···················· | ····················· | ···················· | ···················· | ···················· |
| S | 5642 hCoV-19/Wales/PHWC-48A644/2020\|EPI_ISL_652579\|2020 | ···················· | ···················· | ····················· | ···················· | ···················· | ···················· |
| S | 5643 hCoV-19/Wales/PHWC-48A653/2020\|EPI_ISL_652580\|2020 | ···················· | ···················· | ····················· | ···················· | ···················· | ···················· |
| S | 5644 hCoV-19/Wales/PHWC-48A662/2020\|EPI_ISL_652581\|2020 | ···················· | ···················· | ····················· | ···················· | ···················· | ···················· |
| S | 5645 hCoV-19/Wales/PHWC-48A671/2020\|EPI_ISL_652582\|2020 | ···················· | ···················· | ····················· | ···················· | ···················· | ···················· |
| S | 5646 hCoV-19/Wales/PHWC-48A680/2020\|EPI_ISL_652583\|2020 | ···················· | ···················· | ····················· | ···················· | ···················· | ···················· |
| S | 5647 hCoV-19/Wales/PHWC-48A69F/2020\|EPI_ISL_652584\|2020 | ···················· | ···················· | ····················· | ···················· | ···················· | ···················· |
| S | 5648 hCoV-19/Wales/PHWC-48A6AE/2020\|EPI_ISL_652585\|2020 | ···················· | ···················· | ····················· | ···················· | ···················· | ···················· |
| S | 5649 hCoV-19/Wales/PHWC-48A6BD/2020\|EPI_ISL_652586\|2020 | ···················· | ···················· | ····················· | ···················· | ···················· | ···················· |
| S | 5650 hCoV-19/Wales/PHWC-48A6CC/2020\|EPI_ISL_652587\|2020 | ···················· | ···················· | ····················· | ···················· | ···················· | ···················· |
| S | 5651 hCoV-19/Wales/PHWC-48A6DB/2020\|EPI_ISL_652588\|2020 | ···················· | ···················· | ····················· | ···················· | ···················· | ···················· |
| S | 5652 hCoV-19/Wales/PHWC-48A6EA/2020\|EPI_ISL_652589\|2020 | ···················· | ···················· | ····················· | ···················· | ···················· | ···················· |
| S | 5653 hCoV-19/Wales/PHWC-48A6F9/2020\|EPI_ISL_652590\|2020 | ···················· | ···················· | ····················· | ···················· | ···················· | ···················· |
| S | 5654 hCoV-19/Wales/PHWC-48A705/2020\|EPI_ISL_652591\|2020 | ···················· | ···················· | ····················· | ···················· | ···················· | ···················· |
| S | 5655 hCoV-19/Wales/PHWC-48A714/2020\|EPI_ISL_652592\|2020 | ···················· | ···················· | ····················· | ···················· | ···················· | ···················· |
| S | 5656 hCoV-19/Wales/PHWC-48A723/2020\|EPI_ISL_652593\|2020 | ···················· | ···················· | ····················· | ···················· | ···················· | ···················· |
| S | 5657 hCoV-19/Wales/PHWC-48A732/2020\|EPI_ISL_652594\|2020 | ···················· | ···················· | ····················· | ···················· | ···················· | ···················· |
| S | 5658 hCoV-19/Wales/PHWC-48A741/2020\|EPI_ISL_652595\|2020 | ···················· | ···················· | ····················· | ···················· | ···················· | ···················· |
| S | 5659 hCoV-19/Wales/PHWC-48A750/2020\|EPI_ISL_652596\|2020 | ···················· | ···················· | ····················· | ···················· | ···················· | ···················· |
| S | 5660 hCoV-19/Wales/PHWC-48A77E/2020\|EPI_ISL_652597\|2020 | ···················· | ···················· | ····················· | ···················· | ···················· | ···················· |
| S | 5661 hCoV-19/Wales/PHWC-48A78D/2020\|EPI_ISL_652598\|2020 | ···················· | ···················· | ····················· | ···················· | ···················· | ···················· |
| S | 5662 hCoV-19/Wales/PHWC-48A7BA/2020\|EPI_ISL_652599\|2020 | ···················· | ···················· | ····················· | ···················· | ···················· | ···················· |
| S | 5663 hCoV-19/Wales/PHWC-48A8B7/2020\|EPI_ISL_652610\|2020 | ···················· | ···················· | ····················· | ···················· | ···················· | ···················· |
| S | 5664 hCoV-19/Wales/PHWC-48A8E4/2020\|EPI_ISL_652611\|2020 | ···················· | ···················· | ····················· | ···················· | ···················· | ···················· |
| S | 5665 hCoV-19/Wales/PHWC-48A8F3/2020\|EPI_ISL_652612\|2020 | ···················· | ···················· | ····················· | ···················· | ···················· | ···················· |
| S | 5666 hCoV-19/Wales/PHWC-48A90F/2020\|EPI_ISL_652613\|2020 | ···················· | ···················· | ····················· | ···················· | ···················· | ···················· |
| S | 5667 hCoV-19/Wales/PHWC-48A91E/2020\|EPI_ISL_652614\|2020 | ···················· | ···················· | ····················· | ···················· | ···················· | ···················· |
| S | 5668 hCoV-19/Wales/PHWC-48A93C/2020\|EPI_ISL_652615\|2020 | ···················· | ···················· | ····················· | ···················· | ···················· | ···················· |
| S | 5669 hCoV-19/Wales/PHWC-48A94B/2020\|EPI_ISL_652616\|2020 | ···················· | ···················· | ····················· | ···················· | ···················· | ···················· |
| S | 5670 hCoV-19/Wales/PHWC-48A95A/2020\|EPI_ISL_652617\|2020 | ···················· | ···················· | ····················· | ···················· | ···················· | ···················· |
| S | 5671 hCoV-19/Wales/PHWC-48A987/2020\|EPI_ISL_652618\|2020 | ···················· | ···················· | ····················· | ···················· | ···················· | ···················· |
| S | 5672 hCoV-19/Wales/PHWC-48A9C3/2020\|EPI_ISL_652619\|2020 | ···················· | ···················· | ····················· | ···················· | ···················· | ···················· |
| S | 5673 hCoV-19/England/PORT-2D763B/2020\|EPI_ISL_652620\|20 | ···················· | ···················· | ····················· | ···················· | ···················· | ···················· |
| S | 5674 hCoV-19/England/PORT-2D764A/2020\|EPI_ISL_652621\|20 | ···················· | ···················· | ····················· | ···················· | ······T············· | ···················· |
| S | 5675 hCoV-19/England/PORT-2F5ED5/2020\|EPI_ISL_652622\|20 | ···················· | ···················· | ····················· | ···················· | ···················· | ···················· |
| S | 5676 hCoV-19/England/PORT-2F5EF3/2020\|EPI_ISL_652623\|20 | ···················· | ···················· | ····················· | ···················· | ···················· | ···················· |
| S | 5677 hCoV-19/England/PORT-2F5F5A/2020\|EPI_ISL_652624\|20 | ···················· | ···················· | ····················· | ···················· | ···················· | ···················· |
| S | 5678 hCoV-19/England/PORT-2F5F69/2020\|EPI_ISL_652625\|20 | ···················· | ···················· | ····················· | ···················· | ···················· | ···················· |
| S | 5679 hCoV-19/England/PORT-2F5F87/2020\|EPI_ISL_652626\|20 | ···················· | ···················· | ····················· | ···················· | ···················· | ···················· |
| S | 5680 hCoV-19/England/QEUH-A9BFF4/2020\|EPI_ISL_652627\|20 | ···················· | ···················· | ····················· | ···················· | ···················· | ···················· |
| S | 5681 hCoV-19/Scotland/QEUH-A9C306/2020\|EPI_ISL_652628\|2 | ···················· | ···················· | ····················· | ···················· | ····T··············· | ···················· |
| S | 5682 hCoV-19/Scotland/QEUH-A9C315/2020\|EPI_ISL_652629\|2 | ···················· | ···················· | ····················· | ···················· | ···················· | ···················· |
| S | 5683 hCoV-19/Scotland/QEUH-A9C333/2020\|EPI_ISL_652630\|2 | ···················· | ···················· | ····················· | ···················· | ···················· | ···················· |
| S | 5684 hCoV-19/Scotland/QEUH-A9C342/2020\|EPI_ISL_652631\|2 | ···················· | ···················· | ····················· | ···················· | ······T············· | ···················· |
| S | 5685 hCoV-19/Scotland/QEUH-A9C360/2020\|EPI_ISL_652632\|2 | ···················· | ···················· | ····················· | ···················· | ···················· | ···················· |
| S | 5686 hCoV-19/Scotland/QEUH-A9C37F/2020\|EPI_ISL_652633\|2 | ···················· | ···················· | ····················· | ···················· | ···················· | ···················· |
| S | 5687 hCoV-19/Scotland/QEUH-A9C38E/2020\|EPI_ISL_652634\|2 | ···················· | ···················· | ····················· | ···················· | ···················· | ···················· |
| S | 5688 hCoV-19/Scotland/QEUH-A9C39D/2020\|EPI_ISL_652635\|2 | ···················· | ···················· | ····················· | ···················· | ···················· | ···················· |
| S | 5689 hCoV-19/Scotland/QEUH-A9C3AC/2020\|EPI_ISL_652636\|2 | ···················· | ···················· | ····················· | ···················· | ······T············· | ···················· |
| S | 5690 hCoV-19/Scotland/QEUH-A9C3BB/2020\|EPI_ISL_652637\|2 | ···················· | ···················· | ····················· | ···················· | ···················· | ···················· |
| S | 5691 hCoV-19/Scotland/QEUH-A9C3D9/2020\|EPI_ISL_652638\|2 | ···················· | ···················· | ····················· | ···················· | ······T············· | ···················· |
| S | 5692 hCoV-19/Scotland/QEUH-A9C3F7/2020\|EPI_ISL_652639\|2 | ···················· | ···················· | ····················· | ···················· | ···················· | ···················· |
| S | 5693 hCoV-19/Scotland/QEUH-A9C403/2020\|EPI_ISL_652640\|2 | ···················· | ···················· | ····················· | ···················· | ···················· | ···················· |
| S | 5694 hCoV-19/Scotland/QEUH-A9C412/2020\|EPI_ISL_652641\|2 | ···················· | ···················· | ····················· | ···················· | ···················· | ···················· |

| | | | | | | | | |
|---|---|---|---|---|---|---|---|---|
| S | 5695 | hCoV-19/Scotland/QEUH-A9CE6F/2020\|EPI_ISL_652642\|2 | ···················· | ···················· | ····················· | ···················· | ···················· | ···················· |
| S | 5696 | hCoV-19/England/QEUH-A9CE8D/2020\|EPI_ISL_652643\|20 | ···················· | ···················· | ····················· | ···················· | ···················· | ···················· |
| S | 5697 | hCoV-19/Scotland/QEUH-A9CEBA/2020\|EPI_ISL_652644\|2 | ···················· | ···················· | ····················· | ···················· | ···················· | ···················· |
| S | 5698 | hCoV-19/Scotland/QEUH-A9CEF6/2020\|EPI_ISL_652645\|2 | ···················· | ···················· | ····················· | ···················· | ······T············· | ···················· |
| S | 5699 | hCoV-19/England/QEUH-A9CF02/2020\|EPI_ISL_652646\|20 | ···················· | ···················· | ····················· | ···················· | ···················· | ···················· |
| S | 5700 | hCoV-19/England/QEUH-A9CF3F/2020\|EPI_ISL_652647\|20 | ···················· | ···················· | ····················· | ···················· | ···················· | ···················· |
| S | 5701 | hCoV-19/England/QEUH-A9CF4E/2020\|EPI_ISL_652648\|20 | ···················· | ···················· | ····················· | ···················· | ···················· | ···················· |
| S | 5702 | hCoV-19/Scotland/QEUH-A9D059/2020\|EPI_ISL_652649\|2 | ···················· | ···················· | ····················· | ···················· | ···················· | ···················· |
| S | 5703 | hCoV-19/England/QEUH-A9D068/2020\|EPI_ISL_652650\|20 | ···················· | ···················· | ····················· | ···················· | ···················· | ···················· |
| S | 5704 | hCoV-19/England/QEUH-A9D086/2020\|EPI_ISL_652651\|20 | ···················· | ···················· | ····················· | ···················· | ···················· | ···················· |
| S | 5705 | hCoV-19/Scotland/QEUH-A9D0D1/2020\|EPI_ISL_652652\|2 | ···················· | ···················· | ····················· | ···················· | ···················· | ···················· |
| S | 5706 | hCoV-19/Scotland/QEUH-A9D10B/2020\|EPI_ISL_652653\|2 | ···················· | ···················· | ····················· | ···················· | ···················· | ···················· |
| S | 5707 | hCoV-19/Scotland/QEUH-A9D156/2020\|EPI_ISL_652658\|2 | ···················· | ···················· | ····················· | ···················· | ···················· | ···················· |
| S | 5708 | hCoV-19/England/QEUH-A9D165/2020\|EPI_ISL_652659\|20 | ···················· | ···················· | ····················· | ···················· | ···················· | ···················· |
| S | 5709 | hCoV-19/England/QEUH-A9D9A9/2020\|EPI_ISL_652665\|20 | ···················· | ···················· | ····················· | ···················· | ···················· | ···················· |
| S | 5710 | hCoV-19/England/QEUH-A9D9C7/2020\|EPI_ISL_652666\|20 | ···················· | ···················· | ····················· | ···················· | ···················· | ···················· |
| S | 5711 | hCoV-19/England/QEUH-A9D9D6/2020\|EPI_ISL_652667\|20 | ···················· | ···················· | ····················· | ···················· | ···················· | ···················· |
| S | 5712 | hCoV-19/England/QEUH-A9D9E5/2020\|EPI_ISL_652668\|20 | ···················· | ···················· | ····················· | ···················· | ···················· | ···················· |
| S | 5713 | hCoV-19/England/QEUH-A9DA00/2020\|EPI_ISL_652669\|20 | ···················· | ···················· | ····················· | ···················· | ···················· | ···················· |
| S | 5714 | hCoV-19/England/QEUH-A9DA1F/2020\|EPI_ISL_652670\|20 | ···················· | ···················· | ····················· | ···················· | ···················· | ···················· |
| S | 5715 | hCoV-19/Scotland/QEUH-A9DF98/2020\|EPI_ISL_652671\|2 | ···················· | ···················· | ····················· | ···················· | ···················· | ···················· |
| S | 5716 | hCoV-19/Scotland/QEUH-A9DFB6/2020\|EPI_ISL_652672\|2 | ···················· | ···················· | ····················· | ···················· | ···················· | ···················· |
| S | 5717 | hCoV-19/Scotland/QEUH-A9DFC5/2020\|EPI_ISL_652673\|2 | ···················· | ···················· | ····················· | ···················· | ···················· | ···················· |
| S | 5718 | hCoV-19/Scotland/QEUH-A9DFD4/2020\|EPI_ISL_652674\|2 | ···················· | ···················· | ····················· | ···················· | ···················· | ···················· |
| S | 5719 | hCoV-19/England/QEUH-A9F1BE/2020\|EPI_ISL_652675\|20 | ···················· | ···················· | ····················· | ···················· | ···················· | ···················· |
| S | 5720 | hCoV-19/England/204481497/2020\|EPI_ISL_653951\|2020 | ···················· | ···················· | ····················· | ···················· | ···················· | ···················· |
| S | 5721 | hCoV-19/England/204481499/2020\|EPI_ISL_653952\|2020 | ···················· | ···················· | ····················· | ···················· | ···················· | ···················· |
| S | 5722 | hCoV-19/England/204481950/2020\|EPI_ISL_653953\|2020 | ···················· | ···················· | ····················· | ···················· | ···················· | ···················· |
| S | 5723 | hCoV-19/England/204491001/2020\|EPI_ISL_653954\|2020 | ···················· | ···················· | ····················· | ···················· | ···················· | ···················· |
| S | 5724 | hCoV-19/England/204394042/2020\|EPI_ISL_653958\|2020 | ···················· | ···················· | ····················· | ···················· | ······T············· | ···················· |
| S | 5725 | hCoV-19/England/204394052/2020\|EPI_ISL_653959\|2020 | ···················· | ···················· | ····················· | ···················· | ···················· | ···················· |
| S | 5726 | hCoV-19/England/204400610/2020\|EPI_ISL_653960\|2020 | ···················· | ···················· | ····················· | ···················· | ···················· | ···················· |
| S | 5727 | hCoV-19/England/204400735/2020\|EPI_ISL_653961\|2020 | ···················· | ···················· | ····················· | ···················· | ···················· | ···················· |
| S | 5728 | hCoV-19/England/204420622/2020\|EPI_ISL_653962\|2020 | ···················· | ···················· | ····················· | ···················· | ···················· | ···················· |
| S | 5729 | hCoV-19/England/204420790/2020\|EPI_ISL_653963\|2020 | ···················· | ···················· | ····················· | ···················· | ···················· | ···················· |
| S | 5730 | hCoV-19/England/204420852/2020\|EPI_ISL_653964\|2020 | ···················· | ···················· | ····················· | ···················· | ···················· | ···················· |
| S | 5731 | hCoV-19/England/204421012/2020\|EPI_ISL_653965\|2020 | ···················· | ···················· | ····················· | ···················· | ···················· | ···················· |
| S | 5732 | hCoV-19/England/204421079/2020\|EPI_ISL_653966\|2020 | ···················· | ···················· | ····················· | ···················· | ······T············· | ···················· |
| S | 5733 | hCoV-19/England/204421080/2020\|EPI_ISL_653967\|2020 | ···················· | ···················· | ····················· | ···················· | ···················· | ···················· |
| S | 5734 | hCoV-19/England/204421152/2020\|EPI_ISL_653968\|2020 | ···················· | ···················· | ····················· | ···················· | ···················· | ···················· |
| S | 5735 | hCoV-19/England/204440720/2020\|EPI_ISL_653969\|2020 | ···················· | ···················· | ····················· | ···················· | ···················· | ···················· |
| S | 5736 | hCoV-19/England/204441467/2020\|EPI_ISL_653970\|2020 | ···················· | ···················· | ····················· | ···················· | ···················· | ···················· |
| S | 5737 | hCoV-19/England/204441476/2020\|EPI_ISL_653971\|2020 | ···················· | ···················· | ····················· | ···················· | ···················· | ···················· |
| S | 5739 | hCoV-19/England/204441502/2020\|EPI_ISL_653973\|2020 | ···················· | ·A·················· | ····················· | ···················· | ···················· | ···················· |
| S | 5740 | hCoV-19/England/204441503/2020\|EPI_ISL_653974\|2020 | ···················· | ······A············· | ····················· | ···················· | ···················· | ···················· |
| S | 5741 | hCoV-19/England/204441507/2020\|EPI_ISL_653975\|2020 | ···················· | ·A·················· | ····················· | ···················· | ···················· | ···················· |
| S | 5742 | hCoV-19/England/204460556/2020\|EPI_ISL_653976\|2020 | ···················· | ···················· | ····················· | ···················· | ······T············· | ···················· |
| S | 5743 | hCoV-19/England/204460565/2020\|EPI_ISL_653977\|2020 | ···················· | ···················· | ····················· | ···················· | ···················· | ···················· |
| S | 5744 | hCoV-19/England/204460566/2020\|EPI_ISL_653978\|2020 | ···················· | ···················· | ····················· | ···················· | ···················· | ···················· |
| S | 5745 | hCoV-19/England/204460567/2020\|EPI_ISL_653979\|2020 | ···················· | ···················· | ····················· | ···················· | ···················· | ···················· |
| S | 5746 | hCoV-19/England/204460571/2020\|EPI_ISL_653980\|2020 | ···················· | ···················· | ····················· | ···················· | ···················· | ···················· |
| S | 5747 | hCoV-19/England/204462377/2020\|EPI_ISL_653981\|2020 | ···················· | ···················· | ····················· | ···················· | ···················· | ···················· |
| S | 5748 | hCoV-19/England/204462378/2020\|EPI_ISL_653982\|2020 | ···················· | ···················· | ····················· | ···················· | ···················· | ···················· |
| S | 5749 | hCoV-19/England/204462379/2020\|EPI_ISL_653983\|2020 | ···················· | ···················· | ····················· | ···················· | ···················· | ···················· |
| S | 5750 | hCoV-19/England/204462383/2020\|EPI_ISL_653984\|2020 | ···················· | ···················· | ····················· | ···················· | ···················· | ···················· |
| S | 5751 | hCoV-19/England/204481627/2020\|EPI_ISL_653989\|2020 | ···················· | ···················· | ····················· | ···················· | ···················· | ···················· |
| S | 5752 | hCoV-19/England/204481799/2020\|EPI_ISL_653996\|2020 | ···················· | ···················· | ····················· | ···················· | ······T············· | ···················· |
| S | 5753 | hCoV-19/England/204481811/2020\|EPI_ISL_653997\|2020 | ···················· | ···················· | ····················· | ···················· | ···················· | ···················· |
| S | 5754 | hCoV-19/England/204481881/2020\|EPI_ISL_653998\|2020 | ···················· | ···················· | ····················· | ···················· | ···················· | ···················· |
| S | 5755 | hCoV-19/England/204481882/2020\|EPI_ISL_653999\|2020 | ···················· | ···················· | ····················· | ···················· | ···················· | ···················· |
| S | 5756 | hCoV-19/England/ALDP-B45F43/2020\|EPI_ISL_656100\|20 | ···················· | ···················· | ····················· | ···················· | ···················· | ···················· |
| S | 5757 | hCoV-19/England/ALDP-B46DC0/2020\|EPI_ISL_656101\|20 | ···················· | ···················· | ····················· | ···················· | ···················· | ···················· |
| S | 5758 | hCoV-19/England/ALDP-B46522/2020\|EPI_ISL_656102\|20 | ···················· | ···················· | ····················· | ···················· | ···················· | ···················· |
| S | 5759 | hCoV-19/England/ALDP-B46F24/2020\|EPI_ISL_656103\|20 | ···················· | ···················· | ····················· | ···················· | ···················· | ···················· |
| S | 5760 | hCoV-19/England/ALDP-B462FE/2020\|EPI_ISL_656104\|20 | ···················· | ···················· | ····················· | ···················· | ···················· | ···················· |
| S | 5761 | hCoV-19/England/ALDP-B46364/2020\|EPI_ISL_656105\|20 | ···················· | ···················· | ····················· | ···················· | ···················· | ···················· |
| S | 5762 | hCoV-19/England/ALDP-B4611F/2020\|EPI_ISL_656106\|20 | ···················· | ···················· | ····················· | ···················· | ···················· | ···················· |
| S | 5763 | hCoV-19/England/ALDP-B46786/2020\|EPI_ISL_656107\|20 | ···················· | ···················· | ····················· | ···················· | ···················· | ···················· |
| S | 5764 | hCoV-19/England/ALDP-B46355/2020\|EPI_ISL_656108\|20 | ···················· | ···················· | ····················· | ···················· | ···················· | ···················· |
| S | 5765 | hCoV-19/England/ALDP-B45F07/2020\|EPI_ISL_656109\|20 | ···················· | ···················· | ····················· | ···················· | ···················· | ···················· |
| S | 5766 | hCoV-19/England/ALDP-B463EC/2020\|EPI_ISL_656110\|20 | ···················· | ···················· | ····················· | ···················· | ···················· | ···················· |
| S | 5767 | hCoV-19/England/ALDP-B46838/2020\|EPI_ISL_656111\|20 | ···················· | ···················· | ····················· | ···················· | ···················· | ···················· |
| S | 5768 | hCoV-19/England/ALDP-B45C3D/2020\|EPI_ISL_656112\|20 | ···················· | ···················· | ····················· | ···················· | ···················· | ···················· |
| S | 5769 | hCoV-19/England/ALDP-B465B9/2020\|EPI_ISL_656113\|20 | ···················· | ···················· | ····················· | ···················· | ······T············· | ···················· |
| S | 5770 | hCoV-19/England/ALDP-B469CC/2020\|EPI_ISL_656114\|20 | ···················· | ···················· | ····················· | ···················· | ···················· | ···················· |
| S | 5771 | hCoV-19/England/ALDP-B46328/2020\|EPI_ISL_656115\|20 | ···················· | ···················· | ····················· | ···················· | ···················· | ···················· |
| S | 5772 | hCoV-19/England/ALDP-B45A9D/2020\|EPI_ISL_656116\|20 | ···················· | ···················· | ····················· | ···················· | ···················· | ···················· |
| S | 5773 | hCoV-19/England/ALDP-B464DA/2020\|EPI_ISL_656117\|20 | ···················· | ···················· | ····················· | ···················· | ···················· | ···················· |
| S | 5774 | hCoV-19/England/ALDP-B4649E/2020\|EPI_ISL_656118\|20 | ···················· | ···················· | ····················· | ···················· | ···················· | ···················· |
| S | 5775 | hCoV-19/England/ALDP-B461F1/2020\|EPI_ISL_656119\|20 | ···················· | ···················· | ····················· | ···················· | ···················· | ···················· |

| | | | | | | | | |
|---|---|---|---|---|---|---|---|---|
| S | 5776 hCoV-19/England/ALDP-B46B7B/2020\|EPI_ISL_656120\|20 | ···················· | ···················· | ····················· | ···················· | ···················· | ···················· | |
| S | 5777 hCoV-19/England/ALDP-B46D57/2020\|EPI_ISL_656121\|20 | ···················· | ···················· | ····················· | ···················· | ···················· | ···················· | |
| S | 5778 hCoV-19/England/ALDP-B46610/2020\|EPI_ISL_656122\|20 | ···················· | ···················· | ····················· | ···················· | ···················· | ···················· | |
| S | 5779 hCoV-19/England/ALDP-B46C3C/2020\|EPI_ISL_656123\|20 | ···················· | ···················· | ····················· | ···················· | ···················· | ···················· | |
| S | 5780 hCoV-19/England/ALDP-B46892/2020\|EPI_ISL_656124\|20 | ···················· | ···················· | ····················· | ···················· | ···················· | ···················· | |
| S | 5781 hCoV-19/England/ALDP-B46865/2020\|EPI_ISL_656125\|20 | ···················· | ···················· | ····················· | ···················· | ···················· | ···················· | |
| S | 5782 hCoV-19/England/ALDP-B4630A/2020\|EPI_ISL_656126\|20 | ···················· | ···················· | ····················· | ···················· | ···················· | ···················· | |
| S | 5783 hCoV-19/England/ALDP-B46346/2020\|EPI_ISL_656127\|20 | ···················· | ···················· | ····················· | ···················· | ···················· | ···················· | |
| S | 5784 hCoV-19/England/ALDP-B46777/2020\|EPI_ISL_656128\|20 | ··················T· | ···················· | ····················· | ···················· | ···················· | ···················· | 1 |
| S | 5785 hCoV-19/England/ALDP-B46E81/2020\|EPI_ISL_656129\|20 | ···················· | ···················· | ····················· | ···················· | ···················· | ···················· | |
| S | 5786 hCoV-19/England/ALDP-B45DEF/2020\|EPI_ISL_656130\|20 | ···················· | ···················· | ····················· | ···················· | ···················· | ···················· | |
| S | 5787 hCoV-19/England/ALDP-B46B11/2020\|EPI_ISL_656131\|20 | ···················· | ···················· | ····················· | ···················· | ···················· | ···················· | |
| S | 5788 hCoV-19/England/ALDP-B46F7F/2020\|EPI_ISL_656132\|20 | ···················· | ···················· | ····················· | ···················· | ···················· | ···················· | |
| S | 5789 hCoV-19/England/ALDP-B45C00/2020\|EPI_ISL_656133\|20 | ···················· | ···················· | ····················· | ···················· | ···················· | ···················· | |
| S | 5790 hCoV-19/England/ALDP-B46E18/2020\|EPI_ISL_656134\|20 | ···················· | ···················· | ····················· | ···················· | ···················· | ···················· | |
| S | 5791 hCoV-19/England/ALDP-B47020/2020\|EPI_ISL_656135\|20 | ···················· | ···················· | ····················· | ···················· | ···················· | ···················· | |
| S | 5792 hCoV-19/England/ALDP-B45C4C/2020\|EPI_ISL_656136\|20 | ···················· | ···················· | ····················· | ···················· | ···················· | ···················· | |
| S | 5793 hCoV-19/England/ALDP-B46917/2020\|EPI_ISL_656137\|20 | ···················· | ···················· | ····················· | ···················· | ···················· | ···················· | |
| S | 5794 hCoV-19/England/ALDP-B4671D/2020\|EPI_ISL_656138\|20 | ···················· | ···················· | ····················· | ···················· | ···················· | ···················· | |
| S | 5795 hCoV-19/England/ALDP-B4667A/2020\|EPI_ISL_656139\|20 | ···················· | ···················· | ····················· | ···················· | ···················· | ···················· | |
| S | 5796 hCoV-19/England/MILK-B56659/2020\|EPI_ISL_657406\|20 | ···················· | ···················· | ····················· | ···················· | ···················· | ···················· | |
| S | 5797 hCoV-19/England/MILK-B56343/2020\|EPI_ISL_657410\|20 | ···················· | ···················· | ····················· | ···················· | ···················· | ···················· | |
| S | 5798 hCoV-19/England/MILK-B55915/2020\|EPI_ISL_657411\|20 | ···················· | ···················· | ····················· | ···················· | ···················· | ···················· | |
| S | 5799 hCoV-19/England/CAMC-B532EF/2020\|EPI_ISL_657412\|20 | ···················· | ···················· | ····················· | ···················· | ······T············· | ···················· | |
| S | 5800 hCoV-19/England/MILK-B56219/2020\|EPI_ISL_657413\|20 | ···················· | ·A·················· | ····················· | ···················· | ···················· | ···A················ | |
| S | 5801 hCoV-19/England/MILK-B5571B/2020\|EPI_ISL_657414\|20 | ···················· | ···················· | ····················· | ···················· | ···················· | ···················· | |
| S | 5802 hCoV-19/England/CAMC-B527D2/2020\|EPI_ISL_657415\|20 | ···················· | ···················· | ····················· | ···················· | ···················· | ···················· | |
| S | 5803 hCoV-19/England/MILK-B55854/2020\|EPI_ISL_657416\|20 | ···················· | ···················· | ····················· | ···················· | ···················· | ···················· | |
| S | 5804 hCoV-19/England/MILK-B5639E/2020\|EPI_ISL_657428\|20 | ···················· | ···················· | ····················· | ···················· | ···················· | ···················· | |
| S | 5805 hCoV-19/England/MILK-B55AF4/2020\|EPI_ISL_657429\|20 | ···················· | ···················· | ····················· | ···················· | ···················· | ···················· | |
| S | 5806 hCoV-19/England/MILK-B5598E/2020\|EPI_ISL_657430\|20 | ···················· | ···················· | ····················· | ···················· | ···················· | ···················· | |
| S | 5807 hCoV-19/England/CAMC-B52DB2/2020\|EPI_ISL_657431\|20 | ···················· | ···················· | ····················· | ···················· | ···················· | ···················· | |
| S | 5808 hCoV-19/England/CAMC-B52F61/2020\|EPI_ISL_657432\|20 | ···················· | ···················· | ····················· | ···················· | ······T············· | ···················· | |
| S | 5809 hCoV-19/England/CAMC-B52B03/2020\|EPI_ISL_657433\|20 | ···················· | ···················· | ····················· | ···················· | ···················· | ···················· | |
| S | 5810 hCoV-19/England/CAMC-B52918/2020\|EPI_ISL_657434\|20 | ···················· | ···················· | ····················· | ···················· | ···················· | ···················· | |
| S | 5811 hCoV-19/England/CAMC-B532B2/2020\|EPI_ISL_657436\|20 | ···················· | ···················· | ····················· | ···················· | ···················· | ···················· | |
| S | 5812 hCoV-19/England/CAMC-B52BB8/2020\|EPI_ISL_657437\|20 | ···················· | ···················· | ····················· | ···················· | ···················· | ···················· | |
| S | 5813 hCoV-19/England/MILK-B55757/2020\|EPI_ISL_657438\|20 | ···················· | ···················· | ····················· | ···················· | ···················· | ···················· | |
| S | 5814 hCoV-19/England/MILK-B565D4/2020\|EPI_ISL_657439\|20 | ···················· | ···················· | ····················· | ···················· | ······T············· | ···················· | |
| S | 5815 hCoV-19/England/MILK-B557FD/2020\|EPI_ISL_657440\|20 | ···················· | ·A·················· | ····················· | ···················· | ···················· | ···A················ | |
| S | 5816 hCoV-19/England/MILK-B55845/2020\|EPI_ISL_657441\|20 | ···················· | ···················· | ····················· | ···················· | ···················· | ···················· | |
| S | 5817 hCoV-19/England/MILK-B56149/2020\|EPI_ISL_657443\|20 | ···················· | ···················· | ····················· | ···················· | ···················· | ···················· | |
| S | 5818 hCoV-19/England/CAMC-B52B9A/2020\|EPI_ISL_657444\|20 | ···················· | ···················· | ····················· | ···················· | ······T············· | ···················· | |
| S | 5819 hCoV-19/England/CAMC-B52B30/2020\|EPI_ISL_657445\|20 | ···················· | ···················· | ····················· | ···················· | ···················· | ···················· | |
| S | 5820 hCoV-19/England/MILK-B5564B/2020\|EPI_ISL_657446\|20 | ···················· | ···················· | ····················· | ···················· | ···················· | ···················· | |
| S | 5821 hCoV-19/England/CAMC-B52D2B/2020\|EPI_ISL_657448\|20 | ···················· | ···················· | ····················· | ···················· | ······T············· | ···················· | |
| S | 5822 hCoV-19/England/CAMC-B52DC1/2020\|EPI_ISL_657449\|20 | ···················· | ···················· | ····················· | ···················· | ···················· | ···················· | |
| S | 5823 hCoV-19/England/MILK-B55C1C/2020\|EPI_ISL_657451\|20 | ···················· | ···················· | ····················· | ···················· | ···················· | ···················· | |
| S | 5824 hCoV-19/England/MILK-B56677/2020\|EPI_ISL_657453\|20 | ···················· | ···················· | ····················· | ···················· | ···················· | ···················· | |
| S | 5825 hCoV-19/England/CAMC-B52BF4/2020\|EPI_ISL_657454\|20 | ···················· | ···················· | ····················· | ···················· | ···················· | ···················· | |
| S | 5826 hCoV-19/England/CAMC-B532C1/2020\|EPI_ISL_657456\|20 | ···················· | ···················· | ····················· | ···················· | ······T············· | ···················· | |
| S | 5827 hCoV-19/England/MILK-B555E4/2020\|EPI_ISL_657457\|20 | ···················· | ···················· | ····················· | ···················· | ···················· | ···················· | |
| S | 5828 hCoV-19/England/MILK-B5562D/2020\|EPI_ISL_657459\|20 | ···················· | ···················· | ····················· | ···················· | ···················· | ···················· | |
| S | 5829 hCoV-19/England/CAMC-B528C0/2020\|EPI_ISL_657461\|20 | ···················· | ···················· | ····················· | ···················· | ···················· | ···················· | |
| S | 5830 hCoV-19/England/MILK-B56431/2020\|EPI_ISL_657464\|20 | ···················· | ···················· | ····················· | ···················· | ······T············· | ···················· | |
| S | 5831 hCoV-19/England/CAMC-B52839/2020\|EPI_ISL_657469\|20 | ···················· | ···················· | ····················· | ···················· | ···················· | ···················· | |
| S | 5832 hCoV-19/England/QEUH-B3F38C/2020\|EPI_ISL_658700\|20 | ···················· | ···················· | ····················· | ···················· | ···················· | ···················· | |
| S | 5833 hCoV-19/England/QEUH-B3F4B6/2020\|EPI_ISL_658701\|20 | ···················· | ···················· | ····················· | ···················· | ···················· | ···················· | |
| S | 5834 hCoV-19/England/QEUH-B410DB/2020\|EPI_ISL_658702\|20 | ···················· | ···················· | ····················· | ···················· | ···················· | ···················· | |
| S | 5835 hCoV-19/England/QEUH-B41202/2020\|EPI_ISL_658703\|20 | ···················· | ···················· | ····················· | ···················· | ···················· | ···A················ | |
| S | 5836 hCoV-19/England/QEUH-B3F97B/2020\|EPI_ISL_658704\|20 | ···················· | ···················· | ····················· | ···················· | ···················· | ···················· | |
| S | 5837 hCoV-19/England/QEUH-B4130F/2020\|EPI_ISL_658705\|20 | ···················· | ···················· | ····················· | ···················· | ···················· | ···················· | |
| S | 5838 hCoV-19/England/QEUH-B3F54A/2020\|EPI_ISL_658706\|20 | ···················· | ·A·················· | ····················· | ···················· | ···················· | ···················· | |
| S | 5839 hCoV-19/England/QEUH-B3F28F/2020\|EPI_ISL_658707\|20 | ···················· | ···················· | ····················· | ···················· | ···················· | ···················· | |
| S | 5840 hCoV-19/England/QEUH-B3FBEE/2020\|EPI_ISL_658708\|20 | ···················· | ···················· | ····················· | ···················· | ···················· | ···················· | |
| S | 5841 hCoV-19/England/QEUH-B41572/2020\|EPI_ISL_658709\|20 | ···················· | ···················· | ····················· | ···················· | ···················· | ···················· | |
| S | 5842 hCoV-19/England/QEUH-B41985/2020\|EPI_ISL_658710\|20 | ···················· | ·A·················· | ····················· | ···················· | ···················· | ···················· | |
| S | 5843 hCoV-19/England/QEUH-B411AB/2020\|EPI_ISL_658711\|20 | ···················· | ···················· | ····················· | ···················· | ···················· | ···················· | |
| S | 5844 hCoV-19/England/QEUH-B4119C/2020\|EPI_ISL_658712\|20 | ···················· | ···················· | ····················· | ···················· | ···················· | ···················· | |
| S | 5845 hCoV-19/England/QEUH-B3F53B/2020\|EPI_ISL_658713\|20 | ···················· | ···················· | ····················· | ···················· | ···················· | ···················· | |
| S | 5846 hCoV-19/England/QEUH-B3FCAF/2020\|EPI_ISL_658714\|20 | ···················· | ···················· | ····················· | ···················· | ···················· | ···················· | |
| S | 5847 hCoV-19/England/QEUH-B4176D/2020\|EPI_ISL_658737\|20 | ···················· | ···················· | ····················· | ···················· | ···················· | ···················· | |
| S | 5848 hCoV-19/England/QEUH-B416D9/2020\|EPI_ISL_658738\|20 | ···················· | ···················· | ····················· | ···················· | ···················· | ···················· | |
| S | 5849 hCoV-19/England/QEUH-B3FB0C/2020\|EPI_ISL_658740\|20 | ···················· | ···················· | ····················· | ···················· | ···················· | ···················· | |
| S | 5850 hCoV-19/England/MILK-B39319/2020\|EPI_ISL_658741\|20 | ···················· | ···················· | ····················· | ···················· | ···················· | ···················· | |
| S | 5851 hCoV-19/England/MILK-B38909/2020\|EPI_ISL_658742\|20 | ···················· | ···················· | ····················· | ···················· | ···················· | ···················· | |
| S | 5852 hCoV-19/England/MILK-B39425/2020\|EPI_ISL_658743\|20 | ···················· | ·A·················· | ····················· | ···················· | ···················· | ···A················ | |
| S | 5853 hCoV-19/England/CAMC-B38110/2020\|EPI_ISL_658744\|20 | ···················· | ···················· | ····················· | ···················· | ···················· | ···················· | |
| S | 5854 hCoV-19/England/CAMC-B38031/2020\|EPI_ISL_658745\|20 | ···················· | ···················· | ····················· | ···················· | ···················· | ···················· | |
| S | 5855 hCoV-19/England/MILK-B38927/2020\|EPI_ISL_658746\|20 | ···················· | ···················· | ····················· | ···················· | ···················· | ···················· | |

| | | | | | | | |
|---|---|---|---|---|---|---|---|
| S | 5856 hCoV-19/England/MILK-B38972/2020\|EPI_ISL_658747\|20 | ···················· | ···················· | ···················· | ···················· | ···················· | ···················· |
| S | 5857 hCoV-19/England/CAMC-B37DE0/2020\|EPI_ISL_658759\|20 | ···················· | ···················· | ···················· | ···················· | ···················· | ···················· |
| S | 5858 hCoV-19/England/CAMC-B37E47/2020\|EPI_ISL_658760\|20 | ···················· | ···················· | ···················· | ···················· | ···················· | ···················· |
| S | 5859 hCoV-19/England/MILK-B38945/2020\|EPI_ISL_658761\|20 | ···················· | ···················· | ···················· | ···················· | ···················· | ···················· |
| S | 5860 hCoV-19/England/MILK-B3A64C/2020\|EPI_ISL_658765\|20 | ···················· | ···················· | ···················· | ···················· | ···················· | ···················· |
| S | 5861 hCoV-19/England/CAMC-B37F80/2020\|EPI_ISL_658767\|20 | ···················· | ···················· | ···················· | ···················· | ···················· | ···················· |
| S | 5862 hCoV-19/England/CAMC-B38295/2020\|EPI_ISL_658768\|20 | ···················· | ···················· | ···················· | ···················· | ···················· | ···················· |
| S | 5863 hCoV-19/England/CAMC-B3816B/2020\|EPI_ISL_658769\|20 | ···················· | ·A·················· | ···················· | ···················· | ···················· | ··A················· |
| S | 5864 hCoV-19/England/MILK-B38EBF/2020\|EPI_ISL_658770\|20 | ···················· | ···················· | ···················· | ···················· | ···················· | ···················· |
| S | 5865 hCoV-19/England/MILK-B3A58B/2020\|EPI_ISL_658772\|20 | ···················· | ···················· | ···················· | ···················· | ···················· | ···················· |
| S | 5866 hCoV-19/England/MILK-B3880C/2020\|EPI_ISL_658773\|20 | ···················· | ···················· | ···················· | ···················· | ···················· | ···················· |
| S | 5867 hCoV-19/England/MILK-B38A9D/2020\|EPI_ISL_658774\|20 | ···················· | ···················· | ···················· | ···················· | ···················· | ···················· |
| S | 5868 hCoV-19/England/CAMC-B37ECF/2020\|EPI_ISL_658775\|20 | ···················· | ···················· | ···················· | ···················· | ···················· | ···················· |
| S | 5869 hCoV-19/England/MILK-B38CA6/2020\|EPI_ISL_658776\|20 | ···················· | ···················· | ···················· | ···················· | ···················· | ···················· |
| S | 5870 hCoV-19/England/MILK-B3A600/2020\|EPI_ISL_658777\|20 | ···················· | ···················· | ···················· | ···················· | ···················· | ···················· |
| S | 5871 hCoV-19/England/MILK-B38B4F/2020\|EPI_ISL_658778\|20 | ···················· | ···················· | ···················· | ···················· | ···················· | ···················· |
| S | 5872 hCoV-19/England/MILK-B39328/2020\|EPI_ISL_658779\|20 | ···················· | ···················· | ···················· | ···················· | ···················· | ···················· |
| S | 5873 hCoV-19/England/MILK-B3A56D/2020\|EPI_ISL_658780\|20 | ···················· | ···················· | ···················· | ···················· | ···················· | ···················· |
| S | 5874 hCoV-19/England/CAMC-B3814D/2020\|EPI_ISL_658781\|20 | ···················· | ···················· | ···················· | ···················· | ···················· | ···················· |
| S | 5875 hCoV-19/England/MILK-B383DE/2020\|EPI_ISL_658782\|20 | ···················· | ···················· | ···················· | ···················· | ···················· | ···················· |
| S | 5876 hCoV-19/England/MILK-B38D1C/2020\|EPI_ISL_658783\|20 | ···················· | ···················· | ···················· | ···················· | ···················· | ···················· |
| S | 5877 hCoV-19/England/MILK-B3863F/2020\|EPI_ISL_658784\|20 | ···················· | ···················· | ···················· | ···················· | ···················· | ···················· |
| S | 5878 hCoV-19/England/MILK-B3858D/2020\|EPI_ISL_658785\|20 | ···················· | ···················· | ···················· | ···················· | ···················· | ···················· |
| S | 5879 hCoV-19/England/MILK-B38D76/2020\|EPI_ISL_658786\|20 | ···················· | ···················· | ···················· | ···················· | ···················· | ···················· |
| S | 5880 hCoV-19/England/CAMC-B37D0E/2020\|EPI_ISL_658787\|20 | ···················· | ·A·················· | ···················· | ···················· | ···················· | ··A················· |
| S | 5881 hCoV-19/England/CAMC-B38101/2020\|EPI_ISL_658788\|20 | ···················· | ·A·················· | ···················· | ···················· | ···················· | ··A················· |
| S | 5882 hCoV-19/England/MILK-B3A5B8/2020\|EPI_ISL_658789\|20 | ···················· | ·A·················· | ···················· | ···················· | ···················· | ···················· |
| S | 5883 hCoV-19/England/MILK-B38893/2020\|EPI_ISL_658792\|20 | ···················· | ···················· | ···················· | ···················· | ···················· | ···················· |
| S | 5884 hCoV-19/England/MILK-B3949E/2020\|EPI_ISL_658793\|20 | ···················· | ···················· | ···················· | ···················· | ···················· | ···················· |
| S | 5885 hCoV-19/England/MILK-B38EDD/2020\|EPI_ISL_658796\|20 | ···················· | ···················· | ···················· | ···················· | ···················· | ···················· |
| S | 5886 hCoV-19/England/MILK-B3873C/2020\|EPI_ISL_658797\|20 | ···················· | ·A·················· | ···················· | ···················· | ···················· | ··A················· |
| S | 5887 hCoV-19/England/MILK-B38857/2020\|EPI_ISL_658798\|20 | ···················· | ···················· | ···················· | ···················· | ···················· | ···················· |
| S | 5888 hCoV-19/England/MILK-B38B5E/2020\|EPI_ISL_658799\|20 | ···················· | ···················· | ···················· | ···················· | ···················· | ···················· |
| S | 5889 hCoV-19/Wales/PHWC-48A1CB/2020\|EPI_ISL_652533\|2020 | ···················· | ···················· | ···················· | ···················· | ···················· | ···················· |
| S | 5890 hCoV-19/Wales/PHWC-48A1DA/2020\|EPI_ISL_652534\|2020 | ···················· | ···················· | ···················· | ···················· | ···················· | ···················· |
| S | 5892 hCoV-19/Wales/PHWC-48A1F8/2020\|EPI_ISL_652536\|2020 | ···················· | ···················· | ···················· | ···················· | ···················· | ···················· |
| S | 5893 hCoV-19/Wales/PHWC-48A222/2020\|EPI_ISL_652537\|2020 | ···················· | ···················· | ···················· | ···················· | ···················· | ···················· |
| S | 5894 hCoV-19/Wales/PHWC-48A25F/2020\|EPI_ISL_652538\|2020 | ···················· | ···················· | ···················· | ···················· | ···················· | ···················· |
| S | 5895 hCoV-19/Wales/PHWC-48A28C/2020\|EPI_ISL_652539\|2020 | ···················· | ···················· | ···················· | ···················· | ···················· | ···················· |
| S | 5896 hCoV-19/Wales/PHWC-48A2AA/2020\|EPI_ISL_652540\|2020 | ···················· | ···················· | ···················· | ···················· | ···················· | ···················· |
| S | 5897 hCoV-19/Wales/PHWC-48A2F5/2020\|EPI_ISL_652541\|2020 | ···················· | ···················· | ···················· | ···················· | ···················· | ···················· |
| S | 5898 hCoV-19/Wales/PHWC-48A301/2020\|EPI_ISL_652542\|2020 | ···················· | ···················· | ···················· | ···················· | ···················· | ···················· |
| S | 5899 hCoV-19/Wales/PHWC-48A310/2020\|EPI_ISL_652543\|2020 | ···················· | ···················· | ···················· | ···················· | ···················· | ···················· |
| S | 5900 hCoV-19/Wales/PHWC-48A32F/2020\|EPI_ISL_652544\|2020 | ···················· | ···················· | ···················· | ···················· | ···················· | ···················· |
| S | 5901 hCoV-19/Wales/PHWC-48A34D/2020\|EPI_ISL_652545\|2020 | ···················· | ···················· | ···················· | ···················· | ···················· | ···················· |
| S | 5902 hCoV-19/Wales/PHWC-48A389/2020\|EPI_ISL_652546\|2020 | ···················· | ···················· | ···················· | ···················· | ···················· | ···················· |
| S | 5903 hCoV-19/Wales/PHWC-48A398/2020\|EPI_ISL_652547\|2020 | ···················· | ···················· | ···················· | ···················· | ···················· | ···················· |
| S | 5904 hCoV-19/Wales/PHWC-48A3D4/2020\|EPI_ISL_652550\|2020 | ···················· | ···················· | ···················· | ···················· | ···················· | ···················· |
| S | 5905 hCoV-19/Wales/PHWC-48A3E3/2020\|EPI_ISL_652551\|2020 | ···················· | ···················· | ···················· | ···················· | ···················· | ···················· |
| S | 5906 hCoV-19/Wales/PHWC-48A3F2/2020\|EPI_ISL_652552\|2020 | ···················· | ···················· | ···················· | ···················· | ···················· | ···················· |
| S | 5907 hCoV-19/Wales/PHWC-48A42C/2020\|EPI_ISL_652553\|2020 | ···················· | ···················· | ···················· | ···················· | ···················· | ···················· |
| S | 5908 hCoV-19/Wales/PHWC-48A43B/2020\|EPI_ISL_652554\|2020 | ···················· | ···················· | ···················· | ···················· | ···················· | ···················· |
| S | 5909 hCoV-19/Wales/PHWC-48A459/2020\|EPI_ISL_652555\|2020 | ···················· | ···················· | ···················· | ···················· | ···················· | ···················· |
| S | 5910 hCoV-19/Wales/PHWC-48A468/2020\|EPI_ISL_652556\|2020 | ···················· | ···················· | ···················· | ···················· | ···················· | ···················· |
| S | 5911 hCoV-19/Wales/PHWC-48A486/2020\|EPI_ISL_652557\|2020 | ···················· | ···················· | ···················· | ···················· | ···················· | ···················· |
| S | 5912 hCoV-19/Wales/PHWC-48A495/2020\|EPI_ISL_652558\|2020 | ···················· | ···················· | ···················· | ···················· | ···················· | ···················· |
| S | 5913 hCoV-19/Wales/PHWC-48A4A4/2020\|EPI_ISL_652559\|2020 | ···················· | ···················· | ···················· | ···················· | ···················· | ···················· |
| S | 5914 hCoV-19/Wales/PHWC-48A4C2/2020\|EPI_ISL_652560\|2020 | ···················· | ···················· | ···················· | ···················· | ···················· | ···················· |
| S | 5915 hCoV-19/Wales/PHWC-48A4D1/2020\|EPI_ISL_652561\|2020 | ···················· | ···················· | ···················· | ···················· | ···················· | ···················· |
| S | 5916 hCoV-19/Wales/PHWC-48A4E0/2020\|EPI_ISL_652562\|2020 | ···················· | ···················· | ···················· | ···················· | ···················· | ···················· |
| S | 5917 hCoV-19/Wales/PHWC-48A50B/2020\|EPI_ISL_652563\|2020 | ···················· | ···················· | ···················· | ···················· | ···················· | ···················· |
| S | 5918 hCoV-19/Wales/PHWC-48A51A/2020\|EPI_ISL_652564\|2020 | ···················· | ···················· | ···················· | ···················· | ···················· | ···················· |
| S | 5919 hCoV-19/Wales/PHWC-48A529/2020\|EPI_ISL_652565\|2020 | ···················· | ···················· | ···················· | ···················· | ···················· | ···················· |
| S | 5920 hCoV-19/Wales/PHWC-48A538/2020\|EPI_ISL_652566\|2020 | ···················· | ···················· | ···················· | ···················· | ···················· | ···················· |
| S | 5921 hCoV-19/Wales/PHWC-48A547/2020\|EPI_ISL_652567\|2020 | ···················· | ···················· | ···················· | ···················· | ···················· | ···················· |
| S | 5923 hCoV-19/Wales/PHWC-48A574/2020\|EPI_ISL_652569\|2020 | ···················· | ···················· | ···················· | ···················· | ···················· | ···················· |
| S | 5925 hCoV-19/Wales/PHWC-48A5A1/2020\|EPI_ISL_652571\|2020 | ···················· | ···················· | ···················· | ···················· | ···················· | ···················· |
| S | 5926 hCoV-19/Wales/PHWC-48A5B0/2020\|EPI_ISL_652572\|2020 | ···················· | ···················· | ···················· | ···················· | ···················· | ···················· |
| S | 5927 hCoV-19/Wales/PHWC-48A5CF/2020\|EPI_ISL_652573\|2020 | ···················· | ···················· | ···················· | ···················· | ···················· | ···················· |
| S | 5928 hCoV-19/Wales/PHWC-48A5ED/2020\|EPI_ISL_652574\|2020 | ···················· | ···················· | ···················· | ···················· | ···················· | ···················· |
| S | 5929 hCoV-19/Wales/PHWC-48A608/2020\|EPI_ISL_652575\|2020 | ···················· | ···················· | ···················· | ···················· | ···················· | ···················· |
| S | 5930 hCoV-19/Wales/PHWC-48A617/2020\|EPI_ISL_652576\|2020 | ···················· | ···················· | ···················· | ···················· | ···················· | ···················· |
| S | 5931 hCoV-19/Italy/APU-IZSPB_306PT/2020\|EPI_ISL_653800\| | ···················· | ···················· | ···················· | ···················· | ···················· | ···················· |
| S | 5932 hCoV-19/Italy/APU-IZSPB_307PT/2020\|EPI_ISL_653801\| | ···················· | ···················· | ···················· | ···················· | ···················· | ···················· |
| S | 5933 hCoV-19/Italy/APU-IZSPB_308PT/2020\|EPI_ISL_653802\| | ···················· | ···················· | ···················· | ···················· | ···················· | ···················· |
| S | 5934 hCoV-19/Italy/APU-IZSPB_309PT/2020\|EPI_ISL_653803\| | ···················· | ···················· | ···················· | ···················· | ···················· | ···················· |
| S | 5935 hCoV-19/Italy/APU-IZSPB_310PT/2020\|EPI_ISL_653804\| | ···················· | ···················· | ···················· | ···················· | ···················· | ···················· |
| S | 5936 hCoV-19/Italy/APU-IZSPB_311PT/2020\|EPI_ISL_653805\| | ···················· | ···················· | ···················· | ···················· | ···················· | ···················· |
| S | 5937 hCoV-19/Italy/APU-IZSPB_312PT/2020\|EPI_ISL_653806\| | ···················· | ···················· | ···················· | ···················· | ···················· | ···················· |
| S | 5938 hCoV-19/Italy/APU-IZSPB_313PT/2020\|EPI_ISL_653807\| | ···················· | ···················· | ···················· | ···················· | ···················· | ···················· |

| | | | | | | | | |
|---|---|---|---|---|---|---|---|---|
| S | 5939 hCoV-19/Italy/APU-IZSPB_314PT/2020\|EPI_ISL_653808\| | ···················· | ···················· | ···················· | ···················· | ···················· | ···················· | |
| S | 5940 hCoV-19/Italy/APU-IZSPB_315PT/2020\|EPI_ISL_653809\| | ···················· | ·A·················· | ···················· | ···················· | ···················· | ···················· | |
| S | 5941 hCoV-19/Italy/APU-IZSPB_316PT/2020\|EPI_ISL_653810\| | ···················· | ·A·················· | ···················· | ···················· | ···················· | ···················· | |
| S | 5942 hCoV-19/Italy/APU-IZSPB_317PT/2020\|EPI_ISL_653811\| | ···················· | ·A·················· | ···················· | ···················· | ···················· | ···················· | |
| S | 5943 hCoV-19/Italy/APU-IZSPB_318PT/2020\|EPI_ISL_653812\| | ···················· | ···················· | ···················· | ···················· | ···················· | ···················· | |
| S | 5944 hCoV-19/Italy/APU-IZSPB_319PT/2020\|EPI_ISL_653813\| | ···················· | ···················· | ···················· | ···················· | ···················· | ···················· | |
| S | 5945 hCoV-19/Italy/APU-IZSPB_321PT/2020\|EPI_ISL_653814\| | ···················· | ···················· | ···················· | ···················· | ···················· | ···················· | |
| S | 5946 hCoV-19/Italy/APU-IZSPB_322PT/2020\|EPI_ISL_653815\| | ···················· | ···················· | ···················· | ···················· | ···················· | ···················· | |
| S | 5947 hCoV-19/Italy/APU-IZSPB_323PT/2020\|EPI_ISL_653816\| | ···················· | ···················· | ···················· | ···················· | ···················· | ···················· | |
| S | 5948 hCoV-19/Italy/APU-IZSPB_324PT/2020\|EPI_ISL_653817\| | ···················· | ···················· | ···················· | ···················· | ···················· | ···················· | |
| S | 5949 hCoV-19/Italy/APU-IZSPB_325PT/2020\|EPI_ISL_653818\| | ···················· | ···················· | ···················· | ···················· | ···················· | ···················· | |
| S | 5950 hCoV-19/Italy/APU-IZSPB_326PT/2020\|EPI_ISL_653819\| | ···················· | ···················· | ···················· | ···················· | ······T············· | ···················· | |
| S | 5951 hCoV-19/India/HR-NIBG-TF31/2020\|EPI_ISL_653864\|202 | ···················· | ·A·················· | ···················· | ···················· | ···················· | ···A················ | |
| S | 5952 hCoV-19/India/HR-NIBG-TF32/2020\|EPI_ISL_653865\|202 | ···················· | ···················· | ···················· | ···················· | ···················· | ···················· | |
| S | 5953 hCoV-19/India/HR-NIBG-TF33/2020\|EPI_ISL_653866\|202 | ···················· | ·A·················· | ···················· | ···················· | ···················· | ···A················ | |
| S | 5954 hCoV-19/India/HR-NIBG-TF40/2020\|EPI_ISL_653869\|202 | ···················· | ···················· | ···················· | ···················· | ···················· | ···················· | |
| S | 5955 hCoV-19/India/HR-NIBG-TF44/2020\|EPI_ISL_653871\|202 | ···················· | ·A·················· | ···················· | ···················· | ···················· | ···A················ | |
| S | 5956 hCoV-19/India/HR-NIBG-TF45/2020\|EPI_ISL_653872\|202 | ···················· | ···················· | ···················· | ···················· | ···················· | ···················· | |
| S | 5957 hCoV-19/India/HR-NIBG-TF46/2020\|EPI_ISL_653873\|202 | ···················· | ···················· | ···················· | ···················· | ···················· | ···················· | |
| S | 5958 hCoV-19/India/HR-NIBG-TF47/2020\|EPI_ISL_653874\|202 | ···················· | ···················· | ···················· | ···················· | ···················· | ···················· | |
| S | 5959 hCoV-19/India/HR-NIBG-TF48/2020\|EPI_ISL_653875\|202 | ···················· | ·A·················· | ···················· | ···················· | ···················· | ···A················ | |
| S | 5960 hCoV-19/India/HR-NIBG-TF49/2020\|EPI_ISL_653876\|202 | ···················· | ···················· | ···················· | ···················· | ···················· | ···················· | |
| S | 5961 hCoV-19/India/HR-NIBG-TF50/2020\|EPI_ISL_653877\|202 | ···················· | ·A·················· | ···················· | ···················· | ···················· | ···A················ | |
| S | 5962 hCoV-19/India/HR-NIBG-TF51/2020\|EPI_ISL_653878\|202 | ···················· | ···················· | ···················· | ···················· | ···················· | ···················· | |
| S | 5963 hCoV-19/India/HR-NIBG-TF55/2020\|EPI_ISL_653880\|202 | ···················· | ···················· | ···················· | ···················· | ···················· | ···················· | |
| S | 5964 hCoV-19/India/HR-NIBG-TF56/2020\|EPI_ISL_653881\|202 | ···················· | ···················· | ···················· | ···················· | ···················· | ···················· | |
| S | 5965 hCoV-19/India/HR-NIBG-TF58/2020\|EPI_ISL_653882\|202 | ···················· | ···················· | ···················· | ···················· | ···················· | ···················· | |
| S | 5966 hCoV-19/India/HR-NIBG-TF59/2020\|EPI_ISL_653883\|202 | ···················· | ···················· | ···················· | ···················· | ···················· | ···················· | |
| S | 5967 hCoV-19/India/HR-NIBG-TF61/2020\|EPI_ISL_653885\|202 | ···················· | ···················· | ···················· | ···················· | ···················· | ···················· | |
| S | 5968 hCoV-19/India/HR-NIBG-TF66/2020\|EPI_ISL_653886\|202 | ···················· | ···················· | ···················· | ···················· | ···················· | ···················· | |
| S | 5969 hCoV-19/India/HR-NIBG-TF69/2020\|EPI_ISL_653887\|202 | ···················· | ···················· | ···················· | ···················· | ···················· | ···················· | |
| S | 5970 hCoV-19/India/HR-NIBG-TF70/2020\|EPI_ISL_653888\|202 | ···················· | ·A·················· | ···················· | ···················· | ···················· | ···A················ | |
| S | 5971 hCoV-19/India/HR-NIBG-TF72/2020\|EPI_ISL_653889\|202 | ···················· | ·A·················· | ···················· | ···················· | ···················· | ···A················ | |
| S | 5972 hCoV-19/India/HR-NIBG-TF73/2020\|EPI_ISL_653890\|202 | ···················· | ·A·················· | ···················· | ···················· | ···················· | ···A················ | |
| S | 5973 hCoV-19/India/HR-NIBG-TF74/2020\|EPI_ISL_653891\|202 | ···················· | ···················· | ···················· | ···················· | ···················· | ···················· | |
| S | 5974 hCoV-19/India/HR-NIBG-TF76/2020\|EPI_ISL_653893\|202 | ···················· | ···················· | ···················· | ···················· | ···················· | ···················· | |
| S | 5975 hCoV-19/India/HR-NIBG-TF78/2020\|EPI_ISL_653894\|202 | ···················· | ···················· | ···················· | ···················· | ···················· | ···················· | |
| S | 5976 hCoV-19/India/HR-NIBG-TF79/2020\|EPI_ISL_653895\|202 | ···················· | ···················· | ···················· | ···················· | ···················· | ···················· | |
| S | 5977 hCoV-19/India/HR-NIBG-TF80/2020\|EPI_ISL_653896\|202 | ···················· | ·A·················· | ···················· | ···················· | ···················· | ···A················ | |
| S | 5978 hCoV-19/India/HR-NIBG-TF81/2020\|EPI_ISL_653897\|202 | ···················· | ···················· | ···················· | ···················· | ···················· | ···················· | |
| S | 5979 hCoV-19/India/HR-NIBG-TF82/2020\|EPI_ISL_653898\|202 | ···················· | ···················· | ···················· | ···················· | ···················· | ···················· | |
| S | 5980 hCoV-19/India/HR-NIBG-TF83/2020\|EPI_ISL_653899\|202 | ···················· | ···················· | ···················· | ···················· | ···················· | ···················· | |
| S | 5981 hCoV-19/England/ALDP-B45AE8/2020\|EPI_ISL_656023\|20 | ···················· | ···················· | ···················· | ···················· | ···················· | ···················· | |
| S | 5982 hCoV-19/England/ALDP-B46B20/2020\|EPI_ISL_656024\|20 | ···················· | ···················· | ···················· | ···················· | ···················· | ···················· | |
| S | 5983 hCoV-19/England/ALDP-B4680B/2020\|EPI_ISL_656025\|20 | ··················T· | ···················· | ···················· | ···················· | ···················· | ···················· | 1 |
| S | 5984 hCoV-19/England/ALDP-B46A9C/2020\|EPI_ISL_656026\|20 | ···················· | ···················· | ···················· | ···················· | ···················· | ···················· | |
| S | 5985 hCoV-19/England/ALDP-B459DC/2020\|EPI_ISL_656027\|20 | ···················· | ···················· | ···················· | ···················· | ···················· | ···················· | |
| S | 5986 hCoV-19/England/ALDP-B45ACA/2020\|EPI_ISL_656028\|20 | ···················· | ···················· | ···················· | ···················· | ···················· | ···················· | |
| S | 5987 hCoV-19/England/ALDP-B45C88/2020\|EPI_ISL_656030\|20 | ···················· | ···················· | ···················· | ···················· | ···················· | ···················· | |
| S | 5988 hCoV-19/England/ALDP-B46759/2020\|EPI_ISL_656031\|20 | ···················· | ···················· | ···················· | ···················· | ···················· | ···················· | |
| S | 5989 hCoV-19/England/ALDP-B46391/2020\|EPI_ISL_656032\|20 | ···················· | ···················· | ···················· | ···················· | ···················· | ···················· | |
| S | 5990 hCoV-19/England/ALDP-B45ECE/2020\|EPI_ISL_656033\|20 | ···················· | ···················· | ···················· | ···················· | ···················· | ···················· | |
| S | 5991 hCoV-19/England/ALDP-B462A3/2020\|EPI_ISL_656034\|20 | ···················· | ···················· | ···················· | ···················· | ···················· | ···················· | |
| S | 5992 hCoV-19/England/ALDP-B46ECD/2020\|EPI_ISL_656035\|20 | ···················· | ···················· | ···················· | ···················· | ···················· | ···················· | |
| S | 5993 hCoV-19/England/ALDP-B46DB1/2020\|EPI_ISL_656036\|20 | ···················· | ···················· | ···················· | ···················· | ···················· | ···················· | |
| S | 5994 hCoV-19/England/ALDP-B45FDA/2020\|EPI_ISL_656037\|20 | ···················· | ···················· | ···················· | ···················· | ···················· | ···················· | |
| S | 5995 hCoV-19/England/ALDP-B46EEB/2020\|EPI_ISL_656038\|20 | ···················· | ····A··············· | ···················· | ···················· | ···················· | ···················· | |
| S | 5996 hCoV-19/England/ALDP-B4606D/2020\|EPI_ISL_656039\|20 | ···················· | ···················· | ···················· | ···················· | ···················· | ···················· | |
| S | 5997 hCoV-19/England/ALDP-B46B02/2020\|EPI_ISL_656040\|20 | ···················· | ···················· | ···················· | ···················· | ···················· | ···················· | |
| S | 5998 hCoV-19/England/ALDP-B4614C/2020\|EPI_ISL_656042\|20 | ···················· | ···················· | ···················· | ···················· | ···················· | ···················· | |
| S | 5999 hCoV-19/England/ALDP-B46C2D/2020\|EPI_ISL_656043\|20 | ···················· | ···················· | ···················· | ···················· | ···················· | ···················· | |
| S | 6000 hCoV-19/England/ALDP-B4609A/2020\|EPI_ISL_656046\|20 | ···················· | ···················· | ···················· | ···················· | ···················· | ···················· | |
| S | 6001 hCoV-19/England/ALDP-B472FD/2020\|EPI_ISL_656047\|20 | ···················· | ···················· | ···················· | ···················· | ···················· | ···················· | |
| S | 6002 hCoV-19/England/ALDP-B46C87/2020\|EPI_ISL_656048\|20 | ···················· | ···················· | ···················· | ···················· | ···················· | ···················· | |
| S | 6003 hCoV-19/England/ALDP-B470B7/2020\|EPI_ISL_656049\|20 | ···················· | ·A·················· | ···················· | ···················· | ···················· | ···················· | |
| S | 6004 hCoV-19/England/ALDP-B46FD9/2020\|EPI_ISL_656050\|20 | ···················· | ···················· | ···················· | ···················· | ···················· | ···················· | |
| S | 6005 hCoV-19/England/ALDP-B462B2/2020\|EPI_ISL_656051\|20 | ···················· | ···················· | ···················· | ···················· | ···················· | ···················· | |
| S | 6006 hCoV-19/England/ALDP-B45BB8/2020\|EPI_ISL_656052\|20 | ···················· | ···················· | ···················· | ···················· | ···················· | ···················· | |
| S | 6007 hCoV-19/England/ALDP-B46689/2020\|EPI_ISL_656053\|20 | ···················· | ···················· | ···················· | ···················· | ···················· | ···················· | |
| S | 6008 hCoV-19/England/ALDP-B46E27/2020\|EPI_ISL_656055\|20 | ···················· | ···················· | ···················· | ···················· | ···················· | ···················· | |
| S | 6009 hCoV-19/England/ALDP-B466D4/2020\|EPI_ISL_656056\|20 | ···················· | ···················· | ···················· | ···················· | ···················· | ···················· | |
| S | 6010 hCoV-19/England/ALDP-B460D6/2020\|EPI_ISL_656057\|20 | ···················· | ···················· | ···················· | ···················· | ···················· | ···················· | |
| S | 6011 hCoV-19/England/ALDP-B4699F/2020\|EPI_ISL_656058\|20 | ···················· | ···················· | ···················· | ···················· | ···················· | ···················· | |
| S | 6012 hCoV-19/England/ALDP-B45C79/2020\|EPI_ISL_656059\|20 | ···················· | ···················· | ···················· | ···················· | ···················· | ···················· | |
| S | 6013 hCoV-19/England/ALDP-B45BF4/2020\|EPI_ISL_656060\|20 | ···················· | ···················· | ···················· | ···················· | ···················· | ···················· | |
| S | 6014 hCoV-19/England/ALDP-B461A6/2020\|EPI_ISL_656061\|20 | ···················· | ···················· | ···················· | ···················· | ···················· | ···················· | |
| S | 6015 hCoV-19/England/ALDP-B45A60/2020\|EPI_ISL_656062\|20 | ···················· | ···················· | ···················· | ···················· | ···················· | ···················· | |
| S | 6016 hCoV-19/England/ALDP-B46980/2020\|EPI_ISL_656063\|20 | ···················· | ···················· | ···················· | ···················· | ···················· | ···················· | |
| S | 6017 hCoV-19/England/ALDP-B45EA0/2020\|EPI_ISL_656064\|20 | ···················· | ···················· | ···················· | ···················· | ···················· | ···················· | |
| S | 6018 hCoV-19/England/ALDP-B45F61/2020\|EPI_ISL_656065\|20 | ···················· | ···················· | ···················· | ···················· | ···················· | ···················· | |

| | | | | | | | |
|---|---|---|---|---|---|---|---|
| S | 6019 hCoV-19/England/ALDP-B4622B/2020\|EPI_ISL_656066\|20 | ···················· | ···················· | ····················· | ···················· | ···················· | ···················· |
| S | 6020 hCoV-19/England/ALDP-B46C69/2020\|EPI_ISL_656067\|20 | ···················· | ···················· | ····················· | ···················· | ···················· | ···················· |
| S | 6021 hCoV-19/England/ALDP-B46B6C/2020\|EPI_ISL_656069\|20 | ···················· | ···················· | ····················· | ···················· | ···················· | ···················· |
| S | 6022 hCoV-19/England/ALDP-B46461/2020\|EPI_ISL_656070\|20 | ···················· | ···················· | ····················· | ···················· | ···················· | ···················· |
| S | 6023 hCoV-19/England/ALDP-B4612E/2020\|EPI_ISL_656071\|20 | ···················· | ···················· | ····················· | ···················· | ···················· | ···················· |
| S | 6024 hCoV-19/England/ALDP-B4620D/2020\|EPI_ISL_656072\|20 | ···················· | ···················· | ····················· | ···················· | ···················· | ···················· |
| S | 6025 hCoV-19/England/ALDP-B46F8E/2020\|EPI_ISL_656074\|20 | ···················· | ···················· | ····················· | ···················· | ···················· | ···················· |
| S | 6026 hCoV-19/England/ALDP-B46E90/2020\|EPI_ISL_656075\|20 | ···················· | ···················· | ····················· | ···················· | ···················· | ···················· |
| S | 6027 hCoV-19/England/ALDP-B46DFD/2020\|EPI_ISL_656076\|20 | ···················· | ···················· | ····················· | ···················· | ···················· | ···················· |
| S | 6028 hCoV-19/England/ALDP-B472A2/2020\|EPI_ISL_656077\|20 | ···················· | ···················· | ····················· | ···················· | ···················· | ···················· |
| S | 6029 hCoV-19/England/ALDP-B46BB7/2020\|EPI_ISL_656078\|20 | ···················· | ···················· | ····················· | ···················· | ···················· | ···················· |
| S | 6030 hCoV-19/England/ALDP-B47336/2020\|EPI_ISL_656079\|20 | ···················· | ···················· | ····················· | ···················· | ···················· | ···················· |
| S | 6031 hCoV-19/England/ALDP-B46452/2020\|EPI_ISL_656080\|20 | ···················· | ···················· | ····················· | ···················· | ···················· | ···················· |
| S | 6032 hCoV-19/England/ALDP-B46A14/2020\|EPI_ISL_656081\|20 | ···················· | ···················· | ····················· | ···················· | ···················· | ···················· |
| S | 6033 hCoV-19/England/ALDP-B46F33/2020\|EPI_ISL_656082\|20 | ···················· | ···················· | ····················· | ···················· | ···················· | ···················· |
| S | 6034 hCoV-19/England/ALDP-B46AD8/2020\|EPI_ISL_656084\|20 | ···················· | ···················· | ····················· | ···················· | ···················· | ···················· |
| S | 6035 hCoV-19/England/ALDP-B46030/2020\|EPI_ISL_656085\|20 | ···················· | ···················· | ····················· | ···················· | ···················· | ···················· |
| S | 6036 hCoV-19/England/ALDP-B45E37/2020\|EPI_ISL_656086\|20 | ···················· | ···················· | ····················· | ···················· | ···················· | ···················· |
| S | 6037 hCoV-19/England/ALDP-B4607C/2020\|EPI_ISL_656087\|20 | ···················· | ···················· | ····················· | ···················· | ···················· | ···················· |
| S | 6038 hCoV-19/England/ALDP-B45963/2020\|EPI_ISL_656088\|20 | ···················· | ···················· | ····················· | ···················· | ···················· | ···················· |
| S | 6039 hCoV-19/England/ALDP-B46E63/2020\|EPI_ISL_656089\|20 | ···················· | ···················· | ····················· | ···················· | ···················· | ···················· |
| S | 6040 hCoV-19/England/ALDP-B46188/2020\|EPI_ISL_656091\|20 | ···················· | ···················· | ····················· | ···················· | ···················· | ···················· |
| S | 6041 hCoV-19/England/ALDP-B46470/2020\|EPI_ISL_656092\|20 | ···················· | ···················· | ····················· | ···················· | ···················· | ···················· |
| S | 6042 hCoV-19/England/ALDP-B46D75/2020\|EPI_ISL_656093\|20 | ···················· | ···················· | ····················· | ···················· | ···················· | ···················· |
| S | 6043 hCoV-19/England/ALDP-B46EAF/2020\|EPI_ISL_656094\|20 | ···················· | ···················· | ····················· | ···················· | ···················· | ···················· |
| S | 6044 hCoV-19/England/ALDP-B46CE1/2020\|EPI_ISL_656095\|20 | ···················· | ···················· | ····················· | ···················· | ···················· | ···················· |
| S | 6045 hCoV-19/England/ALDP-B45B4F/2020\|EPI_ISL_656097\|20 | ···················· | ···················· | ····················· | ···················· | ···················· | ···················· |
| S | 6046 hCoV-19/England/ALDP-B45AF7/2020\|EPI_ISL_656098\|20 | ···················· | ···················· | ····················· | ···················· | ···················· | ···················· |
| S | 6047 hCoV-19/England/ALDP-B463FB/2020\|EPI_ISL_656099\|20 | ···················· | ···················· | ····················· | ···················· | ···················· | ···················· |
| S | 6048 hCoV-19/England/QEUH-B40634/2020\|EPI_ISL_657350\|20 | ···················· | ···················· | ····················· | ···················· | ···················· | ···················· |
| S | 6049 hCoV-19/England/QEUH-B4383B/2020\|EPI_ISL_657351\|20 | ···················· | ···················· | ····················· | ···················· | ···················· | ···················· |
| S | 6050 hCoV-19/England/QEUH-B43507/2020\|EPI_ISL_657352\|20 | ···················· | ···················· | ····················· | ···················· | ···················· | ···················· |
| S | 6051 hCoV-19/England/QEUH-B401BB/2020\|EPI_ISL_657353\|20 | ···················· | ·A·················· | ····················· | ···················· | ···················· | ···A················ |
| S | 6052 hCoV-19/England/QEUH-B40090/2020\|EPI_ISL_657354\|20 | ···················· | ···················· | ····················· | ···················· | ···················· | ···················· |
| S | 6053 hCoV-19/England/CAMC-B52AF7/2020\|EPI_ISL_657357\|20 | ···················· | ···················· | ····················· | ···················· | ···················· | ···················· |
| S | 6054 hCoV-19/England/CAMC-B52F07/2020\|EPI_ISL_657358\|20 | ···················· | ···················· | ····················· | ···················· | ···················· | ···················· |
| S | 6055 hCoV-19/England/MILK-B557A2/2020\|EPI_ISL_657359\|20 | ···················· | ···················· | ····················· | ···················· | ···················· | ···················· |
| S | 6056 hCoV-19/England/MILK-B56361/2020\|EPI_ISL_657360\|20 | ···················· | ···················· | ····················· | ···················· | ···················· | ···················· |
| S | 6057 hCoV-19/England/MILK-B556D2/2020\|EPI_ISL_657361\|20 | ···················· | ···················· | ····················· | ···················· | ···················· | ···················· |
| S | 6058 hCoV-19/England/CAMC-B531D3/2020\|EPI_ISL_657362\|20 | ···················· | ···················· | ····················· | ···················· | ······T············· | ···················· |
| S | 6059 hCoV-19/England/MILK-B56176/2020\|EPI_ISL_657365\|20 | ···················· | ···················· | ····················· | ···················· | ···················· | ···················· |
| S | 6060 hCoV-19/England/CAMC-B529BE/2020\|EPI_ISL_657366\|20 | ···················· | ···················· | ····················· | ···················· | ···················· | ···················· |
| S | 6061 hCoV-19/England/MILK-B55A9A/2020\|EPI_ISL_657367\|20 | ···················· | ···················· | ····················· | ···················· | ···················· | ···················· |
| S | 6062 hCoV-19/England/CAMC-B52D3A/2020\|EPI_ISL_657368\|20 | ···················· | ···················· | ····················· | ···················· | ······T············· | ···················· |
| S | 6063 hCoV-19/England/MILK-B5649B/2020\|EPI_ISL_657370\|20 | ···················· | ·A·················· | ····················· | ···················· | ···················· | ···A················ |
| S | 6064 hCoV-19/England/MILK-B558DC/2020\|EPI_ISL_657373\|20 | ···················· | ···················· | ····················· | ···················· | ···················· | ···················· |
| S | 6065 hCoV-19/England/MILK-B558EB/2020\|EPI_ISL_657375\|20 | ···················· | ···················· | ····················· | ···················· | ···················· | ···················· |
| S | 6066 hCoV-19/England/CAMC-B52ACA/2020\|EPI_ISL_657376\|20 | ···················· | ···················· | ····················· | ···················· | ···················· | ···················· |
| S | 6067 hCoV-19/England/MILK-B5657A/2020\|EPI_ISL_657377\|20 | ···················· | ···················· | ····················· | ···················· | ···················· | ···················· |
| S | 6068 hCoV-19/England/MILK-B55678/2020\|EPI_ISL_657381\|20 | ···················· | ···················· | ····················· | ···················· | ···················· | ···················· |
| S | 6069 hCoV-19/England/CAMC-B52B5E/2020\|EPI_ISL_657383\|20 | ···················· | ·A·················· | ····················· | ···················· | ···················· | ···················· |
| S | 6070 hCoV-19/England/MILK-B55A21/2020\|EPI_ISL_657385\|20 | ···················· | ···················· | ····················· | ···················· | ···················· | ···················· |
| S | 6071 hCoV-19/England/MILK-B559E8/2020\|EPI_ISL_657386\|20 | ···················· | ···················· | ····················· | ···················· | ···················· | ···················· |
| S | 6072 hCoV-19/England/CAMC-B52F43/2020\|EPI_ISL_657387\|20 | ···················· | ···················· | ····················· | ···················· | ···················· | ···················· |
| S | 6073 hCoV-19/England/CAMC-B5321C/2020\|EPI_ISL_657388\|20 | ···················· | ···················· | ····················· | ···················· | ···················· | ···················· |
| S | 6074 hCoV-19/England/CAMC-B52F70/2020\|EPI_ISL_657389\|20 | ···················· | ···················· | ····················· | ···················· | ···················· | ···················· |
| S | 6075 hCoV-19/England/MILK-B55AE5/2020\|EPI_ISL_657390\|20 | ···················· | ···················· | ····················· | ···················· | ···················· | ···················· |
| S | 6076 hCoV-19/England/MILK-B558CD/2020\|EPI_ISL_657392\|20 | ···················· | ···················· | ····················· | ···················· | ···················· | ···················· |
| S | 6077 hCoV-19/England/MILK-B55A30/2020\|EPI_ISL_657393\|20 | ···················· | ···················· | ····················· | ···················· | ···················· | ···················· |
| S | 6078 hCoV-19/England/CAMC-B53267/2020\|EPI_ISL_657394\|20 | ···················· | ···················· | ····················· | ···················· | ···················· | ···················· |
| S | 6079 hCoV-19/England/CAMC-B52A42/2020\|EPI_ISL_657395\|20 | ········T··········· | ···················· | ····················· | ···················· | ···················· | ···················· |
| S | 6080 hCoV-19/England/MILK-B56291/2020\|EPI_ISL_657396\|20 | ···················· | ·A·················· | ····················· | ···················· | ···················· | ···A················ |
| S | 6081 hCoV-19/England/CAMC-B529FA/2020\|EPI_ISL_657397\|20 | ···················· | ···················· | ····················· | ···················· | ····T·T············· | ···················· |
| S | 6082 hCoV-19/England/MILK-B565C5/2020\|EPI_ISL_657398\|20 | ···················· | ···················· | ····················· | ···················· | ···················· | ···················· |
| S | 6083 hCoV-19/England/CAMC-B52EDD/2020\|EPI_ISL_657399\|20 | ···················· | ···················· | ····················· | ···················· | ···················· | ···················· |
| S | 6084 hCoV-19/England/MILK-B55836/2020\|EPI_ISL_657401\|20 | ···················· | ···················· | ····················· | ···················· | ···················· | ···················· |
| S | 6085 hCoV-19/England/CAMC-B52CC4/2020\|EPI_ISL_657402\|20 | ···················· | ···················· | ····················· | ···················· | ···················· | ···················· |
| S | 6086 hCoV-19/England/CAMC-B52BD6/2020\|EPI_ISL_657403\|20 | ···················· | ···················· | ····················· | ···················· | ···················· | ···················· |
| S | 6087 hCoV-19/England/CAMC-B532A3/2020\|EPI_ISL_657404\|20 | ···················· | ···················· | ····················· | ···················· | ···················· | ···················· |
| S | 6088 hCoV-19/England/MILK-B55B6A/2020\|EPI_ISL_657405\|20 | ···················· | ···················· | ····················· | ···················· | ···················· | ···················· |
| S | 6089 hCoV-19/England/CAMC-B52884/2020\|EPI_ISL_657407\|20 | ···················· | ···················· | ····················· | ···················· | ···················· | ···················· |
| S | 6090 hCoV-19/England/MILK-B5661D/2020\|EPI_ISL_657408\|20 | ···················· | ·A·················· | ····················· | ···················· | ···················· | ···A················ |
| S | 6091 hCoV-19/England/MILK-B55827/2020\|EPI_ISL_657417\|20 | ···················· | ···················· | ····················· | ···················· | ···················· | ···················· |
| S | 6092 hCoV-19/England/MILK-B56440/2020\|EPI_ISL_657419\|20 | ···················· | ···················· | ····················· | ···················· | ···················· | ···················· |
| S | 6093 hCoV-19/England/CAMC-B5323A/2020\|EPI_ISL_657423\|20 | ···················· | ···················· | ····················· | ···················· | ···················· | ···················· |
| S | 6094 hCoV-19/England/MILK-B55784/2020\|EPI_ISL_657424\|20 | ···················· | ···················· | ····················· | ···················· | ···················· | ···················· |
| S | 6095 hCoV-19/England/MILK-B563E9/2020\|EPI_ISL_657425\|20 | ···················· | ···················· | ····················· | ···················· | ···················· | ···················· |
| S | 6096 hCoV-19/England/CAMC-B52C00/2020\|EPI_ISL_657427\|20 | ···················· | ···················· | ····················· | ···················· | ···················· | ···················· |
| S | 6097 hCoV-19/England/QEUH-B3FB39/2020\|EPI_ISL_658680\|20 | ···················· | ···················· | ····················· | ···················· | ···················· | ···················· |
| S | 6098 hCoV-19/England/QEUH-B412D5/2020\|EPI_ISL_658682\|20 | ···················· | ···················· | ····················· | ···················· | ···················· | ···················· |

| | | | | | | | |
|---|---|---|---|---|---|---|---|
| S | 6099 hCoV-19/England/QEUH-B3F771/2020\|EPI_ISL_658683\|20 | .................... | .A.................. | .................... | .................... | .................... | .................... |
| S | 6100 hCoV-19/England/QEUH-B410EA/2020\|EPI_ISL_658684\|20 | .................... | .................... | .................... | .................... | .................... | .................... |
| S | 6101 hCoV-19/England/QEUH-B3FD33/2020\|EPI_ISL_658685\|20 | .................... | .................... | .................... | .................... | .................... | .................... |
| S | 6102 hCoV-19/England/QEUH-B3F2F8/2020\|EPI_ISL_658688\|20 | .................... | .................... | .................... | .................... | .................... | .................... |
| S | 6103 hCoV-19/England/QEUH-B3F261/2020\|EPI_ISL_658689\|20 | .................... | .................... | .................... | .................... | .................... | .................... |
| S | 6104 hCoV-19/England/QEUH-B41439/2020\|EPI_ISL_658691\|20 | .................... | .................... | .................... | .................... | .................... | .................... |
| S | 6105 hCoV-19/England/QEUH-B4117E/2020\|EPI_ISL_658692\|20 | .................... | .................... | .................... | .................... | .................... | .................... |
| S | 6106 hCoV-19/England/QEUH-B41624/2020\|EPI_ISL_658693\|20 | .................... | .................... | .................... | .................... | .................... | .................... |
| S | 6107 hCoV-19/England/QEUH-B413A5/2020\|EPI_ISL_658694\|20 | .................... | .................... | .................... | .................... | .................... | .................... |
| S | 6108 hCoV-19/England/QEUH-B41B16/2020\|EPI_ISL_658695\|20 | .................... | .................... | .................... | .................... | .................... | .................... |
| S | 6109 hCoV-19/England/QEUH-B4124E/2020\|EPI_ISL_658696\|20 | .................... | .................... | .................... | .................... | .................... | .................... |
| S | 6110 hCoV-19/England/QEUH-B3F5A4/2020\|EPI_ISL_658697\|20 | .................... | .................... | .................... | .................... | .................... | .................... |
| S | 6111 hCoV-19/England/QEUH-B3F823/2020\|EPI_ISL_658698\|20 | .................... | .................... | .................... | .................... | .................... | .................... |
| S | 6112 hCoV-19/England/QEUH-B40F93/2020\|EPI_ISL_658699\|20 | .................... | .................... | .................... | .................... | .................... | .................... |
| S | 6113 hCoV-19/England/QEUH-B40F39/2020\|EPI_ISL_658715\|20 | .................... | .................... | .................... | .................... | .................... | .................... |
| S | 6114 hCoV-19/England/QEUH-B3F304/2020\|EPI_ISL_658717\|20 | .................... | .................... | .................... | .................... | .................... | .................... |
| S | 6115 hCoV-19/England/QEUH-B3F4D4/2020\|EPI_ISL_658719\|20 | .................... | .................... | .................... | .................... | .................... | .................... |
| S | 6116 hCoV-19/England/QEUH-B3F45C/2020\|EPI_ISL_658720\|20 | .................... | .................... | .................... | .................... | .................... | .................... |
| S | 6117 hCoV-19/England/QEUH-B418F1/2020\|EPI_ISL_658721\|20 | .................... | .................... | .................... | .................... | .................... | .................... |
| S | 6118 hCoV-19/England/QEUH-B3F6A1/2020\|EPI_ISL_658723\|20 | .................... | .A.................. | .................... | .................... | .................... | .................... |
| S | 6119 hCoV-19/England/QEUH-B3FBDF/2020\|EPI_ISL_658724\|20 | .................... | .................... | .................... | .................... | .................... | .................... |
| S | 6120 hCoV-19/England/QEUH-B3F8F6/2020\|EPI_ISL_658725\|20 | .................... | .................... | .................... | .................... | .................... | .................... |
| S | 6121 hCoV-19/England/QEUH-B418D3/2020\|EPI_ISL_658726\|20 | .................... | .................... | .................... | .................... | .................... | .................... |
| S | 6122 hCoV-19/England/QEUH-B3F5E0/2020\|EPI_ISL_658727\|20 | .................... | .................... | .................... | .................... | .................... | .................... |
| S | 6123 hCoV-19/England/QEUH-B41536/2020\|EPI_ISL_658731\|20 | .................... | .................... | .................... | .................... | .................... | .................... |
| S | 6124 hCoV-19/England/QEUH-B4125D/2020\|EPI_ISL_658732\|20 | .................... | .................... | .................... | .................... | .................... | .................... |
| S | 6125 hCoV-19/England/QEUH-B415EB/2020\|EPI_ISL_658733\|20 | .................... | .................... | .................... | .................... | .................... | .................... |
| S | 6126 hCoV-19/England/QEUH-B41475/2020\|EPI_ISL_658734\|20 | .................... | .................... | .................... | .................... | .................... | .................... |
| S | 6127 hCoV-19/England/QEUH-B41ABF/2020\|EPI_ISL_658735\|20 | .................... | .................... | .................... | .................... | .................... | .................... |
| S | 6128 hCoV-19/England/QEUH-B3F8AB/2020\|EPI_ISL_658736\|20 | .................... | .................... | .................... | .................... | .................... | .................... |
| S | 6129 hCoV-19/England/MILK-B384BD/2020\|EPI_ISL_658751\|20 | .................... | .................... | .................... | .................... | .................... | .................... |
| S | 6130 hCoV-19/England/CAMC-B3812F/2020\|EPI_ISL_658752\|20 | .................... | .A.................. | .................... | .................... | .................... | ...A................ |
| S | 6131 hCoV-19/England/CAMC-B380D7/2020\|EPI_ISL_658753\|20 | .................... | .................... | .................... | .................... | .................... | .................... |
| S | 6132 hCoV-19/England/MILK-B3A55E/2020\|EPI_ISL_658754\|20 | .................... | .................... | .................... | .................... | .................... | .................... |
| S | 6133 hCoV-19/England/MILK-B3A61F/2020\|EPI_ISL_658755\|20 | .................... | .................... | .................... | .................... | .................... | .................... |
| S | 6134 hCoV-19/England/CAMC-B37F62/2020\|EPI_ISL_658756\|20 | .................... | .................... | .................... | .................... | .................... | .................... |
| S | 6135 hCoV-19/England/MILK-B39179/2020\|EPI_ISL_658757\|20 | .................... | .................... | .................... | .................... | .................... | .................... |
| S | 6136 hCoV-19/England/MILK-B3867B/2020\|EPI_ISL_658758\|20 | .................... | .................... | .................... | .................... | .................... | .................... |
| S | 6137 hCoV-19/USA/WI-GMF-27798/2020\|EPI_ISL_660992\|2020- | .................... | .................... | .................... | .................... | .................... | .................... |
| S | 6138 hCoV-19/USA/WI-GMF-27860/2020\|EPI_ISL_660993\|2020- | .................... | .................... | .................... | .................... | .................... | .................... |
| S | 6139 hCoV-19/USA/WI-GMF-27861/2020\|EPI_ISL_660994\|2020- | .................... | .................... | .................... | .................... | .................... | .................... |
| S | 6140 hCoV-19/USA/WI-GMF-27869/2020\|EPI_ISL_660995\|2020- | .................... | .A.................. | .................... | .................... | .................... | .................... |
| S | 6141 hCoV-19/USA/WI-GMF-27942/2020\|EPI_ISL_660996\|2020- | .................... | .................... | .................... | .................... | .................... | .................... |
| S | 6142 hCoV-19/USA/WI-GMF-27987/2020\|EPI_ISL_660997\|2020- | .................... | .................... | .................... | .................... | .................... | .................... |
| S | 6143 hCoV-19/USA/WI-GMF-27988/2020\|EPI_ISL_660998\|2020- | .................... | .................... | .................... | .................... | .................... | .................... |
| S | 6144 hCoV-19/USA/WI-GMF-28004/2020\|EPI_ISL_660999\|2020- | .................... | .................... | .................... | .................... | .................... | .................... |
| S | 6145 hCoV-19/Wales/PHWC-489B28/2020\|EPI_ISL_652456\|2020 | .................... | .................... | .................... | .................... | .................... | .................... |
| S | 6146 hCoV-19/Wales/PHWC-489B37/2020\|EPI_ISL_652457\|2020 | .................... | .................... | .................... | .................... | .................... | .................... |
| S | 6147 hCoV-19/Wales/PHWC-489B46/2020\|EPI_ISL_652458\|2020 | .................... | .................... | .................... | .................... | .................... | .................... |
| S | 6148 hCoV-19/Wales/PHWC-489B55/2020\|EPI_ISL_652459\|2020 | .................... | .................... | .................... | .................... | .................... | .................... |
| S | 6149 hCoV-19/Wales/PHWC-489B73/2020\|EPI_ISL_652460\|2020 | .................... | .................... | .................... | .................... | .................... | .................... |
| S | 6150 hCoV-19/Wales/PHWC-489B82/2020\|EPI_ISL_652461\|2020 | .................... | .................... | .................... | .................... | .................... | .................... |
| S | 6151 hCoV-19/Wales/PHWC-489B91/2020\|EPI_ISL_652462\|2020 | .................... | .................... | .................... | .................... | .................... | .................... |
| S | 6152 hCoV-19/Wales/PHWC-489BA0/2020\|EPI_ISL_652463\|2020 | .................... | .A.................. | .................... | .................... | .................... | .................... |
| S | 6153 hCoV-19/Wales/PHWC-489BBF/2020\|EPI_ISL_652464\|2020 | .................... | .................... | .................... | .................... | .................... | .................... |
| S | 6154 hCoV-19/Wales/PHWC-489BCE/2020\|EPI_ISL_652465\|2020 | .................... | .................... | .................... | .................... | .................... | .................... |
| S | 6155 hCoV-19/Wales/PHWC-489BDD/2020\|EPI_ISL_652466\|2020 | .................... | .................... | .................... | .................... | .................... | .................... |
| S | 6156 hCoV-19/Wales/PHWC-489BEC/2020\|EPI_ISL_652467\|2020 | .................... | .................... | .................... | .................... | .................... | .................... |
| S | 6157 hCoV-19/Wales/PHWC-489BFB/2020\|EPI_ISL_652468\|2020 | .................... | .................... | .................... | .................... | .................... | .................... |
| S | 6158 hCoV-19/Wales/PHWC-489C16/2020\|EPI_ISL_652469\|2020 | .................... | .................... | .................... | .................... | .................... | .................... |
| S | 6159 hCoV-19/Wales/PHWC-489C25/2020\|EPI_ISL_652470\|2020 | .................... | .................... | .................... | .................... | .................... | .................... |
| S | 6160 hCoV-19/Wales/PHWC-489C34/2020\|EPI_ISL_652471\|2020 | .................... | .................... | .................... | .................... | .................... | .................... |
| S | 6161 hCoV-19/Wales/PHWC-489C43/2020\|EPI_ISL_652472\|2020 | .................... | .................... | .................... | .................... | .................... | .................... |
| S | 6162 hCoV-19/Wales/PHWC-489C52/2020\|EPI_ISL_652473\|2020 | .................... | .................... | .................... | .................... | .................... | .................... |
| S | 6163 hCoV-19/Wales/PHWC-489C70/2020\|EPI_ISL_652474\|2020 | .................... | .................... | .................... | .................... | .................... | .................... |
| S | 6164 hCoV-19/Wales/PHWC-489C8F/2020\|EPI_ISL_652475\|2020 | .................... | .................... | .................... | .................... | .................... | .................... |
| S | 6165 hCoV-19/Wales/PHWC-489C9E/2020\|EPI_ISL_652476\|2020 | .................... | .................... | .................... | .................... | .................... | .................... |
| S | 6166 hCoV-19/Wales/PHWC-489CAD/2020\|EPI_ISL_652477\|2020 | .................... | .................... | .................... | .................... | .................... | .................... |
| S | 6167 hCoV-19/Wales/PHWC-489CBC/2020\|EPI_ISL_652478\|2020 | .................... | .................... | .................... | .................... | .................... | .................... |
| S | 6168 hCoV-19/Wales/PHWC-489CCB/2020\|EPI_ISL_652479\|2020 | .................... | .................... | .................... | .................... | .................... | .................... |
| S | 6169 hCoV-19/Wales/PHWC-489CDA/2020\|EPI_ISL_652480\|2020 | .................... | .................... | .................... | .................... | .................... | .................... |
| S | 6170 hCoV-19/Wales/PHWC-489CE9/2020\|EPI_ISL_652481\|2020 | .................... | .................... | .................... | .................... | .................... | .................... |
| S | 6171 hCoV-19/Wales/PHWC-489CF8/2020\|EPI_ISL_652482\|2020 | .................... | .................... | .................... | .................... | .................... | .................... |
| S | 6172 hCoV-19/Wales/PHWC-489D04/2020\|EPI_ISL_652483\|2020 | .................... | .................... | .................... | .................... | .................... | .................... |
| S | 6173 hCoV-19/Wales/PHWC-489D22/2020\|EPI_ISL_652484\|2020 | .................... | .................... | .................... | .................... | .................... | .................... |
| S | 6174 hCoV-19/Wales/PHWC-489D31/2020\|EPI_ISL_652485\|2020 | .................... | .................... | .................... | .................... | .................... | .................... |
| S | 6175 hCoV-19/Wales/PHWC-489D40/2020\|EPI_ISL_652486\|2020 | .................... | .................... | .................... | .................... | .................... | .................... |
| S | 6176 hCoV-19/Wales/PHWC-489D5F/2020\|EPI_ISL_652487\|2020 | .................... | .................... | .................... | .................... | .................... | .................... |
| S | 6177 hCoV-19/Wales/PHWC-489D6E/2020\|EPI_ISL_652488\|2020 | .................... | .................... | .................... | .................... | .................... | .................... |
| S | 6178 hCoV-19/Wales/PHWC-489D8C/2020\|EPI_ISL_652489\|2020 | .................... | .................... | .................... | .................... | .................... | .................... |

| | | | | | | | |
|---|---|---|---|---|---|---|---|
| S | 6179 hCoV-19/Wales/PHWC-489DAA/2020\|EPI_ISL_652490\|2020 | ···················· | ···················· | ····················· | ···················· | ···················· | ···················· |
| S | 6180 hCoV-19/Wales/PHWC-489DB9/2020\|EPI_ISL_652491\|2020 | ···················· | ···················· | ····················· | ···················· | ···················· | ···················· |
| S | 6181 hCoV-19/Wales/PHWC-489DD7/2020\|EPI_ISL_652492\|2020 | ···················· | ···················· | ····················· | ···················· | ···················· | ···················· |
| S | 6182 hCoV-19/Wales/PHWC-489DE6/2020\|EPI_ISL_652493\|2020 | ···················· | ···················· | ····················· | ···················· | ···················· | ···················· |
| S | 6183 hCoV-19/Wales/PHWC-489DF5/2020\|EPI_ISL_652494\|2020 | ···················· | ···················· | ····················· | ···················· | ···················· | ···················· |
| S | 6184 hCoV-19/Wales/PHWC-489E10/2020\|EPI_ISL_652495\|2020 | ···················· | ···················· | ····················· | ···················· | ···················· | ···················· |
| S | 6185 hCoV-19/Wales/PHWC-489E2F/2020\|EPI_ISL_652496\|2020 | ···················· | ···················· | ····················· | ···················· | ···················· | ···················· |
| S | 6186 hCoV-19/Wales/PHWC-489E3E/2020\|EPI_ISL_652497\|2020 | ···················· | ···················· | ····················· | ···················· | ···················· | ···················· |
| S | 6187 hCoV-19/Wales/PHWC-489E4D/2020\|EPI_ISL_652498\|2020 | ···················· | ···················· | ····················· | ···················· | ···················· | ···················· |
| S | 6188 hCoV-19/Wales/PHWC-489E5C/2020\|EPI_ISL_652499\|2020 | ···················· | ···················· | ····················· | ···················· | ···················· | ···················· |
| S | 6190 hCoV-19/Wales/PHWC-489E89/2020\|EPI_ISL_652501\|2020 | ···················· | ···················· | ····················· | ···················· | ···················· | ···················· |
| S | 6191 hCoV-19/Wales/PHWC-489E98/2020\|EPI_ISL_652502\|2020 | ···················· | ···················· | ····················· | ···················· | ···················· | ···················· |
| S | 6192 hCoV-19/Wales/PHWC-489EC5/2020\|EPI_ISL_652503\|2020 | ···················· | ···················· | ····················· | ···················· | ···················· | ···················· |
| S | 6193 hCoV-19/Wales/PHWC-489ED4/2020\|EPI_ISL_652504\|2020 | ···················· | ···················· | ····················· | ···················· | ···················· | ···················· |
| S | 6194 hCoV-19/Wales/PHWC-489EE3/2020\|EPI_ISL_652505\|2020 | ···················· | ···················· | ····················· | ···················· | ···················· | ···················· |
| S | 6195 hCoV-19/Wales/PHWC-489F3B/2020\|EPI_ISL_652506\|2020 | ···················· | ···················· | ····················· | ···················· | ···················· | ···················· |
| S | 6196 hCoV-19/Wales/PHWC-489F4A/2020\|EPI_ISL_652507\|2020 | ···················· | ···················· | ····················· | ···················· | ···················· | ···················· |
| S | 6197 hCoV-19/Wales/PHWC-489F68/2020\|EPI_ISL_652508\|2020 | ···················· | ···················· | ····················· | ···················· | ···················· | ···················· |
| S | 6198 hCoV-19/Wales/PHWC-489F77/2020\|EPI_ISL_652509\|2020 | ···················· | ···················· | ····················· | ···················· | ···················· | ···················· |
| S | 6199 hCoV-19/Wales/PHWC-489F86/2020\|EPI_ISL_652510\|2020 | ···················· | ···················· | ····················· | ···················· | ···················· | ···················· |
| S | 6200 hCoV-19/Wales/PHWC-489FA4/2020\|EPI_ISL_652511\|2020 | ···················· | ···················· | ····················· | ···················· | ···················· | ···················· |
| S | 6201 hCoV-19/Wales/PHWC-489FB3/2020\|EPI_ISL_652512\|2020 | ···················· | ···················· | ····················· | ···················· | ···················· | ···················· |
| S | 6202 hCoV-19/Wales/PHWC-489FC2/2020\|EPI_ISL_652513\|2020 | ···················· | ···················· | ····················· | ···················· | ···················· | ···················· |
| S | 6203 hCoV-19/Wales/PHWC-489FD1/2020\|EPI_ISL_652514\|2020 | ···················· | ···················· | ····················· | ···················· | ···················· | ···················· |
| S | 6204 hCoV-19/Wales/PHWC-489FE0/2020\|EPI_ISL_652515\|2020 | ···················· | ···················· | ····················· | ···················· | ···················· | ···················· |
| S | 6205 hCoV-19/Wales/PHWC-489FFF/2020\|EPI_ISL_652516\|2020 | ···················· | ···················· | ····················· | ···················· | ···················· | ···················· |
| S | 6206 hCoV-19/Wales/PHWC-48A055/2020\|EPI_ISL_652517\|2020 | ···················· | ···················· | ····················· | ···················· | ···················· | ···················· |
| S | 6207 hCoV-19/Wales/PHWC-48A064/2020\|EPI_ISL_652518\|2020 | ···················· | ·A·················· | ····················· | ···················· | ···················· | ··A················· |
| S | 6208 hCoV-19/Wales/PHWC-48A082/2020\|EPI_ISL_652519\|2020 | ···················· | ·A·················· | ····················· | ···················· | ···················· | ···················· |
| S | 6209 hCoV-19/Wales/PHWC-48A0A0/2020\|EPI_ISL_652520\|2020 | ···················· | ···················· | ····················· | ···················· | ···················· | ···················· |
| S | 6210 hCoV-19/Wales/PHWC-48A0BF/2020\|EPI_ISL_652521\|2020 | ···················· | ···················· | ····················· | ···················· | ···················· | ···················· |
| S | 6211 hCoV-19/Wales/PHWC-48A0DD/2020\|EPI_ISL_652522\|2020 | ···················· | ···················· | ····················· | ···················· | ···················· | ···················· |
| S | 6212 hCoV-19/Wales/PHWC-48A0FB/2020\|EPI_ISL_652523\|2020 | ···················· | ···················· | ····················· | ···················· | ···················· | ···················· |
| S | 6213 hCoV-19/Wales/PHWC-48A107/2020\|EPI_ISL_652524\|2020 | ···················· | ···················· | ····················· | ···················· | ···················· | ···················· |
| S | 6214 hCoV-19/Wales/PHWC-48A116/2020\|EPI_ISL_652525\|2020 | ···················· | ···················· | ····················· | ···················· | ···················· | ···················· |
| S | 6215 hCoV-19/Wales/PHWC-48A125/2020\|EPI_ISL_652526\|2020 | ···················· | ···················· | ····················· | ···················· | ···················· | ···················· |
| S | 6216 hCoV-19/Wales/PHWC-48A152/2020\|EPI_ISL_652527\|2020 | ···················· | ···················· | ····················· | ···················· | ···················· | ···················· |
| S | 6217 hCoV-19/Wales/PHWC-48A161/2020\|EPI_ISL_652528\|2020 | ···················· | ···················· | ····················· | ···················· | ···················· | ···················· |
| S | 6218 hCoV-19/Wales/PHWC-48A170/2020\|EPI_ISL_652529\|2020 | ···················· | ···················· | ····················· | ···················· | ···················· | ···················· |
| S | 6219 hCoV-19/Wales/PHWC-48A18F/2020\|EPI_ISL_652530\|2020 | ···················· | ···················· | ····················· | ···················· | ···················· | ···················· |
| S | 6220 hCoV-19/Wales/PHWC-48A19E/2020\|EPI_ISL_652531\|2020 | ···················· | ···················· | ····················· | ···················· | ···················· | ···················· |
| S | 6221 hCoV-19/Wales/PHWC-48A1BC/2020\|EPI_ISL_652532\|2020 | ···················· | ···················· | ····················· | ···················· | ···················· | ···················· |
| S | 6222 hCoV-19/Wales/PHWC-48A3B6/2020\|EPI_ISL_652548\|2020 | ···················· | ···················· | ····················· | ···················· | ···················· | ···················· |
| S | 6223 hCoV-19/Wales/PHWC-48A3C5/2020\|EPI_ISL_652549\|2020 | ···················· | ···················· | ····················· | ···················· | ···················· | ···················· |
| S | 6224 hCoV-19/Italy/APU-IZSPB_293PT/2020\|EPI_ISL_653787\| | ···················· | ···················· | ····················· | ···················· | ···················· | ···················· |
| S | 6225 hCoV-19/Italy/APU-IZSPB_294PT/2020\|EPI_ISL_653788\| | ···················· | ···················· | ····················· | ···················· | ···················· | ···················· |
| S | 6226 hCoV-19/Italy/APU-IZSPB_295PT/2020\|EPI_ISL_653789\| | ···················· | ···················· | ····················· | ···················· | ···················· | ···················· |
| S | 6227 hCoV-19/Italy/APU-IZSPB_296PT/2020\|EPI_ISL_653790\| | ···················· | ···················· | ····················· | ···················· | ···················· | ···················· |
| S | 6228 hCoV-19/Italy/APU-IZSPB_297PT/2020\|EPI_ISL_653791\| | ···················· | ···················· | ····················· | ···················· | ···················· | ···················· |
| S | 6229 hCoV-19/Italy/BAS-IZSPB_298PT/2020\|EPI_ISL_653792\| | ···················· | ···················· | ····················· | ···················· | ···················· | ···················· |
| S | 6230 hCoV-19/Italy/APU-IZSPB_299PT/2020\|EPI_ISL_653793\| | ···················· | ···················· | ····················· | ···················· | ···················· | ···················· |
| S | 6231 hCoV-19/Italy/APU-IZSPB_300PT/2020\|EPI_ISL_653794\| | ···················· | ···················· | ····················· | ···················· | ···················· | ···················· |
| S | 6232 hCoV-19/Italy/APU-IZSPB_301PT/2020\|EPI_ISL_653795\| | ···················· | ···················· | ····················· | ···················· | ···················· | ···················· |
| S | 6233 hCoV-19/Italy/APU-IZSPB_302PT/2020\|EPI_ISL_653796\| | ···················· | ···················· | ····················· | ···················· | ···················· | ···················· |
| S | 6234 hCoV-19/Italy/APU-IZSPB_303PT/2020\|EPI_ISL_653797\| | ···················· | ···················· | ····················· | ···················· | ···················· | ···················· |
| S | 6235 hCoV-19/Italy/APU-IZSPB_304PT/2020\|EPI_ISL_653798\| | ···················· | ···················· | ····················· | ···················· | ···················· | ···················· |
| S | 6236 hCoV-19/Italy/APU-IZSPB_305PT/2020\|EPI_ISL_653799\| | ···················· | ···················· | ····················· | ···················· | ···················· | ···················· |
| S | 6237 hCoV-19/Italy/APU-IZSPB_327PT/2020\|EPI_ISL_653820\| | ···················· | ···················· | ····················· | ···················· | ···················· | ···················· |
| S | 6238 hCoV-19/Italy/APU-IZSPB_328PT/2020\|EPI_ISL_653821\| | ···················· | ···················· | ····················· | ···················· | ···················· | ···················· |
| S | 6239 hCoV-19/Italy/APU-IZSPB_329PT/2020\|EPI_ISL_653822\| | ···················· | ···················· | ····················· | ···················· | ······T············· | ···················· |
| S | 6240 hCoV-19/Italy/APU-IZSPB_171PT/2020\|EPI_ISL_653823\| | ···················· | ···················· | ····················· | ···················· | ···················· | ···················· |
| S | 6241 hCoV-19/India/WB-NIBG-IK49/2020\|EPI_ISL_653826\|202 | ···················· | ···················· | ····················· | ···················· | ···················· | ···················· |
| S | 6242 hCoV-19/India/WB-NIBG-IK76/2020\|EPI_ISL_653827\|202 | ···················· | ···················· | ····················· | ···················· | ··A················· | ···················· |
| S | 6243 hCoV-19/India/WB-NIBG-IK84/2020\|EPI_ISL_653828\|202 | ···················· | ···················· | ····················· | ···················· | ···················· | ···················· |
| S | 6244 hCoV-19/India/WB-NIBG-IK87/2020\|EPI_ISL_653829\|202 | ···················· | ···················· | ····················· | ···················· | ···················· | ···················· |
| S | 6245 hCoV-19/India/WB-NIBG-IK94/2020\|EPI_ISL_653830\|202 | ···················· | ···················· | ····················· | ···················· | ···················· | ···················· |
| S | 6246 hCoV-19/India/WB-NIBG-IK98/2020\|EPI_ISL_653831\|202 | ···················· | ···················· | ····················· | ···················· | ···················· | ···················· |
| S | 6247 hCoV-19/India/WB-NIBG-IK100/2020\|EPI_ISL_653832\|20 | ···················· | ···················· | ····················· | ···················· | ···················· | ···················· |
| S | 6248 hCoV-19/India/DL-NIBG-MN2/2020\|EPI_ISL_653833\|2020 | ···················· | ·A·················· | ····················· | ···················· | ···················· | ··A················· |
| S | 6249 hCoV-19/India/DL-NIBG-MN3/2020\|EPI_ISL_653834\|2020 | ···················· | ·A·················· | ····················· | ···················· | ···················· | ··A················· |
| S | 6250 hCoV-19/India/DL-NIBG-MN6/2020\|EPI_ISL_653835\|2020 | ···················· | ·A·················· | ····················· | ···················· | ···················· | ··A················· |
| S | 6251 hCoV-19/India/DL-NIBG-MN7/2020\|EPI_ISL_653836\|2020 | ···················· | ···················· | ····················· | ···················· | ···················· | ···················· |
| S | 6252 hCoV-19/India/DL-NIBG-MN9/2020\|EPI_ISL_653837\|2020 | ···················· | ·A·················· | ····················· | ···················· | ···················· | ··A················· |
| S | 6253 hCoV-19/India/DL-NIBG-MN12/2020\|EPI_ISL_653838\|202 | ···················· | ·A·················· | ····················· | ···················· | ···················· | ··A················· |
| S | 6254 hCoV-19/India/DL-NIBG-MN13/2020\|EPI_ISL_653839\|202 | ···················· | ·············A······ | ····················· | ···················· | ···················· | ···················· |
| S | 6255 hCoV-19/India/DL-NIBG-MN14/2020\|EPI_ISL_653840\|202 | ···················· | ·············A······ | ····················· | ···················· | ···················· | ···················· |
| S | 6256 hCoV-19/India/DL-NIBG-MN15/2020\|EPI_ISL_653841\|202 | ···················· | ···················· | ····················· | ···················· | ···················· | ···················· |
| S | 6257 hCoV-19/India/DL-NIBG-MN16/2020\|EPI_ISL_653842\|202 | ···················· | ·A·················· | ····················· | ···················· | ···················· | ··A················· |
| S | 6258 hCoV-19/India/DL-NIBG-MN17/2020\|EPI_ISL_653843\|202 | ···················· | ·A·················· | ····················· | ···················· | ···················· | ··A················· |
| S | 6259 hCoV-19/India/DL-NIBG-MN18/2020\|EPI_ISL_653844\|202 | ···················· | ·A·················· | ····················· | ···················· | ···················· | ··A················· |

| | | | | | | | | |
|---|---|---|---|---|---|---|---|---|
| S | 6260 | hCoV-19/India/DL-NIBG-MN19/2020\|EPI_ISL_653845\|202 | ···················· | ·A·················· | ···················· | ···················· | ···················· | ··A················· |
| S | 6261 | hCoV-19/India/DL-NIBG-MN20/2020\|EPI_ISL_653846\|202 | ···················· | ·A·················· | ···················· | ···················· | ···················· | ··A················· |
| S | 6262 | hCoV-19/India/DL-NIBG-MN21/2020\|EPI_ISL_653847\|202 | ···················· | ·A·················· | ···················· | ···················· | ···················· | ··A················· |
| S | 6263 | hCoV-19/India/DL-NIBG-MN22/2020\|EPI_ISL_653848\|202 | ···················· | ·A·················· | ···················· | ···················· | ···················· | ··A················· |
| S | 6264 | hCoV-19/India/DL-NIBG-MN23/2020\|EPI_ISL_653849\|202 | ···················· | ···················· | ···················· | ···················· | ···················· | ···················· |
| S | 6265 | hCoV-19/India/DL-NIBG-MN24/2020\|EPI_ISL_653850\|202 | ···················· | ·A·················· | ···················· | ···················· | ···················· | ··A················· |
| S | 6266 | hCoV-19/India/DL-NIBG-MN25/2020\|EPI_ISL_653851\|202 | ···················· | ·A·················· | ···················· | ···················· | ···················· | ··A················· |
| S | 6267 | hCoV-19/India/DL-NIBG-MN27/2020\|EPI_ISL_653852\|202 | ···················· | ·A·················· | ···················· | ···················· | ···················· | ··A················· |
| S | 6268 | hCoV-19/India/DL-NIBG-MN28/2020\|EPI_ISL_653853\|202 | ···················· | ·A·················· | ···················· | ···················· | ···················· | ··A················· |
| S | 6269 | hCoV-19/India/DL-NIBG-MN30/2020\|EPI_ISL_653854\|202 | ···················· | ·A·················· | ···················· | ···················· | ···················· | ··A················· |
| S | 6270 | hCoV-19/India/DL-NIBG-MN31/2020\|EPI_ISL_653855\|202 | ···················· | ·A·················· | ···················· | ···················· | ···················· | ··A················· |
| S | 6271 | hCoV-19/India/DL-NIBG-MN33/2020\|EPI_ISL_653856\|202 | ···················· | ·A·················· | ···················· | ···················· | ···················· | ··A················· |
| S | 6272 | hCoV-19/India/DL-NIBG-MN34/2020\|EPI_ISL_653857\|202 | ···················· | ·A·················· | ···················· | ···················· | ···················· | ··A················· |
| S | 6273 | hCoV-19/India/DL-NIBG-MN35/2020\|EPI_ISL_653858\|202 | ···················· | ·A·················· | ···················· | ···················· | ···················· | ··A················· |
| S | 6274 | hCoV-19/India/HR-NIBG-TF5/2020\|EPI_ISL_653859\|2020 | ···················· | ·A·················· | ···················· | ···················· | ···················· | ··A················· |
| S | 6275 | hCoV-19/India/HR-NIBG-TF11/2020\|EPI_ISL_653860\|202 | ···················· | ·A·················· | ···················· | ···················· | ···················· | ··A················· |
| S | 6276 | hCoV-19/India/HR-NIBG-TF18/2020\|EPI_ISL_653861\|202 | ···················· | ·A·················· | ···················· | ···················· | ···················· | ··A················· |
| S | 6277 | hCoV-19/India/HR-NIBG-TF29/2020\|EPI_ISL_653863\|202 | ···················· | ·A·················· | ···················· | ···················· | ···················· | ··A················· |
| S | 6278 | hCoV-19/India/DL-NIBG-TF52/2020\|EPI_ISL_653879\|202 | ···················· | ·A·················· | ···················· | ···················· | ···················· | ··A················· |
| S | 6279 | hCoV-19/England/ALDP-B45B5E/2020\|EPI_ISL_656000\|20 | ···················· | ···················· | ···················· | ···················· | ···················· | ···················· |
| S | 6280 | hCoV-19/England/ALDP-B46C96/2020\|EPI_ISL_656001\|20 | ···················· | ···················· | ···················· | ···················· | ···················· | ···················· |
| S | 6281 | hCoV-19/England/ALDP-B46AAB/2020\|EPI_ISL_656002\|20 | ···················· | ···················· | ···················· | ···················· | ···················· | ···················· |
| S | 6282 | hCoV-19/England/ALDP-B45CB5/2020\|EPI_ISL_656003\|20 | ···················· | ···················· | ···················· | ···················· | ···················· | ···················· |
| S | 6283 | hCoV-19/England/ALDP-B4673B/2020\|EPI_ISL_656004\|20 | ···················· | ···················· | ···················· | ···················· | ···················· | ···················· |
| S | 6284 | hCoV-19/England/ALDP-B46935/2020\|EPI_ISL_656005\|20 | ···················· | ···················· | ···················· | ···················· | ···················· | ···················· |
| S | 6285 | hCoV-19/England/ALDP-B467A4/2020\|EPI_ISL_656006\|20 | ···················· | ···················· | ···················· | ···················· | ···················· | ···················· |
| S | 6286 | hCoV-19/England/ALDP-B45B7C/2020\|EPI_ISL_656007\|20 | ···················· | ···················· | ···················· | ···················· | ···················· | ···················· |
| S | 6287 | hCoV-19/England/ALDP-B4648F/2020\|EPI_ISL_656008\|20 | ···················· | ···················· | ···················· | ···················· | ···················· | ···················· |
| S | 6288 | hCoV-19/England/ALDP-B46953/2020\|EPI_ISL_656009\|20 | ···················· | ···················· | ···················· | ···················· | ···················· | ···················· |
| S | 6289 | hCoV-19/England/ALDP-B45E91/2020\|EPI_ISL_656010\|20 | ···················· | ···················· | ···················· | ···················· | ···················· | ···················· |
| S | 6290 | hCoV-19/England/ALDP-B45D49/2020\|EPI_ISL_656011\|20 | ···················· | ···················· | ···················· | ···················· | ···················· | ···················· |
| S | 6291 | hCoV-19/England/ALDP-B46294/2020\|EPI_ISL_656012\|20 | ···················· | ···················· | ···················· | ···················· | ···················· | ···················· |
| S | 6292 | hCoV-19/England/ALDP-B47354/2020\|EPI_ISL_656013\|20 | ···················· | ···················· | ···················· | ···················· | ···················· | ···················· |
| S | 6293 | hCoV-19/England/ALDP-B4659B/2020\|EPI_ISL_656014\|20 | ···················· | ···················· | ···················· | ···················· | ···················· | ···················· |
| S | 6294 | hCoV-19/England/ALDP-B45FF8/2020\|EPI_ISL_656015\|20 | ···················· | ···················· | ···················· | ···················· | ···················· | ···················· |
| S | 6295 | hCoV-19/England/ALDP-B46425/2020\|EPI_ISL_656017\|20 | ···················· | ···················· | ···················· | ···················· | ···················· | ···················· |
| S | 6296 | hCoV-19/England/ALDP-B4621C/2020\|EPI_ISL_656018\|20 | ···················· | ···················· | ···················· | ···················· | ···················· | ···················· |
| S | 6297 | hCoV-19/England/ALDP-B46A23/2020\|EPI_ISL_656019\|20 | ···················· | ···················· | ···················· | ···················· | ···················· | ···················· |
| S | 6298 | hCoV-19/England/QEUH-B42438/2020\|EPI_ISL_657310\|20 | ···················· | ···················· | ···················· | ···················· | ···················· | ···················· |
| S | 6299 | hCoV-19/England/QEUH-B43886/2020\|EPI_ISL_657314\|20 | ···················· | ···················· | ···················· | ···················· | ···················· | ···················· |
| S | 6300 | hCoV-19/England/QEUH-B421F5/2020\|EPI_ISL_657315\|20 | ···················· | ···················· | ···················· | ···················· | ···················· | ···················· |
| S | 6301 | hCoV-19/England/QEUH-B43552/2020\|EPI_ISL_657316\|20 | ···················· | ···················· | ···················· | ···················· | ···················· | ···················· |
| S | 6302 | hCoV-19/England/QEUH-B423C2/2020\|EPI_ISL_657317\|20 | ···················· | ···················· | ···················· | ···················· | ···················· | ···················· |
| S | 6303 | hCoV-19/England/QEUH-B402B8/2020\|EPI_ISL_657329\|20 | ···················· | ···················· | ···················· | ···················· | ···················· | ···················· |
| S | 6304 | hCoV-19/England/QEUH-B43358/2020\|EPI_ISL_657330\|20 | ···················· | ···················· | ···················· | ···················· | ···················· | ···················· |
| S | 6305 | hCoV-19/England/QEUH-B404A3/2020\|EPI_ISL_657332\|20 | ···················· | ···················· | ···················· | ···················· | ···················· | ···················· |
| S | 6306 | hCoV-19/England/QEUH-B404B2/2020\|EPI_ISL_657333\|20 | ···················· | ···················· | ···················· | ···················· | ···················· | ···················· |
| S | 6307 | hCoV-19/England/QEUH-B43604/2020\|EPI_ISL_657334\|20 | ···················· | ···················· | ···················· | ···················· | ···················· | ···················· |
| S | 6308 | hCoV-19/England/QEUH-B43394/2020\|EPI_ISL_657335\|20 | ···················· | ···················· | ···················· | ···················· | ···················· | ···················· |
| S | 6309 | hCoV-19/England/QEUH-B4230E/2020\|EPI_ISL_657336\|20 | ···················· | ···················· | ···················· | ···················· | ···················· | ···················· |
| S | 6310 | hCoV-19/England/QEUH-B4359E/2020\|EPI_ISL_657337\|20 | ···················· | ·A·················· | ···················· | ···················· | ···················· | ··A················· |
| S | 6311 | hCoV-19/England/QEUH-B406BC/2020\|EPI_ISL_657338\|20 | ···················· | ···················· | ···················· | ···················· | ···················· | ···················· |
| S | 6312 | hCoV-19/England/QEUH-B4084D/2020\|EPI_ISL_657339\|20 | ···················· | ···················· | ···················· | ···················· | ···················· | ···················· |
| S | 6313 | hCoV-19/England/QEUH-B40494/2020\|EPI_ISL_657340\|20 | ···················· | ···················· | ···················· | ···················· | ···················· | ···················· |
| S | 6314 | hCoV-19/England/QEUH-B405DD/2020\|EPI_ISL_657342\|20 | ···················· | ···················· | ···················· | ···················· | ···················· | ···················· |
| S | 6315 | hCoV-19/England/QEUH-B40564/2020\|EPI_ISL_657343\|20 | ···················· | ···················· | ···················· | ···················· | ···················· | ···················· |
| S | 6316 | hCoV-19/England/QEUH-B4027C/2020\|EPI_ISL_657345\|20 | ···················· | ···················· | ···················· | ···················· | ···················· | ···················· |
| S | 6317 | hCoV-19/England/QEUH-B434A0/2020\|EPI_ISL_657346\|20 | ···················· | ···················· | ···················· | ···················· | ···················· | ···················· |
| S | 6318 | hCoV-19/England/QEUH-B40968/2020\|EPI_ISL_657347\|20 | ···················· | ···················· | ···················· | ···················· | ···················· | ···················· |
| S | 6319 | hCoV-19/England/QEUH-B3FE4F/2020\|EPI_ISL_657348\|20 | ···················· | ···················· | ···················· | ···················· | ···················· | ···················· |
| S | 6320 | hCoV-19/England/QEUH-B4240B/2020\|EPI_ISL_657349\|20 | ···················· | ···················· | ···················· | ···················· | ···················· | ···················· |
| S | 6321 | hCoV-19/England/QEUH-B3F243/2020\|EPI_ISL_658640\|20 | ···················· | ···················· | ···················· | ···················· | ···················· | ···················· |
| S | 6322 | hCoV-19/England/QEUH-B41378/2020\|EPI_ISL_658642\|20 | ···················· | ······A············· | ···················· | ···················· | ···················· | ···················· |
| S | 6323 | hCoV-19/England/QEUH-B3FBFD/2020\|EPI_ISL_658645\|20 | ···················· | ···················· | ···················· | ···················· | ···················· | ···················· |
| S | 6324 | hCoV-19/England/QEUH-B41A91/2020\|EPI_ISL_658646\|20 | ···················· | ·A·················· | ···················· | ···················· | ···················· | ···················· |
| S | 6325 | hCoV-19/England/QEUH-B4135A/2020\|EPI_ISL_658647\|20 | ···················· | ···················· | ···················· | ···················· | ···················· | ···················· |
| S | 6326 | hCoV-19/England/QEUH-B41105/2020\|EPI_ISL_658648\|20 | ···················· | ···················· | ···················· | ···················· | ···················· | ···················· |
| S | 6327 | hCoV-19/England/QEUH-B41071/2020\|EPI_ISL_658662\|20 | ···················· | ···················· | ···················· | ···················· | ···················· | ···················· |
| S | 6328 | hCoV-19/England/QEUH-B3F4E3/2020\|EPI_ISL_658663\|20 | ···················· | ···················· | ···················· | ···················· | ···················· | ···················· |
| S | 6329 | hCoV-19/England/QEUH-B4140C/2020\|EPI_ISL_658664\|20 | ···················· | ···················· | ···················· | ···················· | ···················· | ···················· |
| S | 6330 | hCoV-19/England/QEUH-B3FCDC/2020\|EPI_ISL_658665\|20 | ···················· | ···················· | ···················· | ···················· | ···················· | ···················· |
| S | 6331 | hCoV-19/England/QEUH-B41493/2020\|EPI_ISL_658667\|20 | ···················· | ···················· | ···················· | ···················· | ···················· | ···················· |
| S | 6332 | hCoV-19/England/QEUH-B4167F/2020\|EPI_ISL_658669\|20 | ···················· | ···················· | ···················· | ···················· | ···················· | ···················· |
| S | 6333 | hCoV-19/England/QEUH-B411D8/2020\|EPI_ISL_658670\|20 | ···················· | ···················· | ···················· | ···················· | ···················· | ···················· |
| S | 6334 | hCoV-19/England/QEUH-B3F7AE/2020\|EPI_ISL_658671\|20 | ···················· | ···················· | ···················· | ···················· | ···················· | ···················· |
| S | 6335 | hCoV-19/England/QEUH-B40FDF/2020\|EPI_ISL_658672\|20 | ···················· | ···················· | ···················· | ···················· | ···················· | ···················· |
| S | 6336 | hCoV-19/England/QEUH-B4116F/2020\|EPI_ISL_658673\|20 | ···················· | ···················· | ···················· | ···················· | ···················· | ···················· |
| S | 6337 | hCoV-19/England/QEUH-B3FA69/2020\|EPI_ISL_658674\|20 | ···················· | ···················· | ···················· | ···················· | ···················· | ···················· |
| S | 6338 | hCoV-19/England/QEUH-B41615/2020\|EPI_ISL_658675\|20 | ···················· | ···················· | ···················· | ···················· | ···················· | ···················· |
| S | 6339 | hCoV-19/England/QEUH-B410AE/2020\|EPI_ISL_658676\|20 | ···················· | ···················· | ···················· | ···················· | ···················· | ···················· |

| | | | | | | | |
|---|---|---|---|---|---|---|---|
| S | 6340 hCoV-19/England/QEUH-B3F2DA/2020\|EPI_ISL_658677\|20 | ···················· | ···················· | ····················· | ···················· | ···················· | ···················· |
| S | 6341 hCoV-19/England/QEUH-B3FB84/2020\|EPI_ISL_658678\|20 | ···················· | ···················· | ····················· | ···················· | ···················· | ···················· |
| S | 6342 hCoV-19/England/QEUH-B3FC18/2020\|EPI_ISL_658679\|20 | ···················· | ···················· | ····················· | ···················· | ···················· | ···················· |
| S | 6343 hCoV-19/England/QEUH-B2CE72/2020\|EPI_ISL_659900\|20 | ···················· | ···················· | ····················· | ···················· | ······T············· | ···················· |
| S | 6344 hCoV-19/England/QEUH-B2EDCE/2020\|EPI_ISL_659902\|20 | ···················· | ···················· | ····················· | ···················· | ···················· | ···················· |
| S | 6345 hCoV-19/England/QEUH-B2EB00/2020\|EPI_ISL_659907\|20 | ···················· | ···················· | ····················· | ···················· | ···················· | ···················· |
| S | 6346 hCoV-19/England/QEUH-B2DFD8/2020\|EPI_ISL_659908\|20 | ···················· | ·A·················· | ····················· | ···················· | ···················· | ···················· |
| S | 6347 hCoV-19/England/QEUH-B2D5F4/2020\|EPI_ISL_659910\|20 | ···················· | ·A·················· | ····················· | ···················· | ···················· | ···················· |
| S | 6348 hCoV-19/England/QEUH-B2E8BE/2020\|EPI_ISL_659912\|20 | ···················· | ···················· | ····················· | ···················· | ···················· | ···················· |
| S | 6349 hCoV-19/England/QEUH-B3042E/2020\|EPI_ISL_659913\|20 | ···················· | ···················· | ····················· | ···················· | ···················· | ···················· |
| S | 6350 hCoV-19/England/QEUH-B30BA1/2020\|EPI_ISL_659915\|20 | ···················· | ···················· | ····················· | ···················· | ···················· | ···················· |
| S | 6351 hCoV-19/England/QEUH-B2EE61/2020\|EPI_ISL_659917\|20 | ···················· | ···················· | ····················· | ···················· | ···················· | ···················· |
| S | 6352 hCoV-19/England/QEUH-B2CDC0/2020\|EPI_ISL_659918\|20 | ···················· | ···················· | ····················· | ···················· | ···················· | ···················· |
| S | 6353 hCoV-19/England/QEUH-B2DFC9/2020\|EPI_ISL_659919\|20 | ···················· | ···················· | ····················· | ···················· | ···················· | ···················· |
| S | 6354 hCoV-19/England/QEUH-B2DF41/2020\|EPI_ISL_659920\|20 | ···················· | ···················· | ····················· | ···················· | ···················· | ···················· |
| S | 6355 hCoV-19/England/QEUH-B2E915/2020\|EPI_ISL_659921\|20 | ···················· | ···················· | ····················· | ···················· | ···················· | ···················· |
| S | 6356 hCoV-19/England/QEUH-B2CD1B/2020\|EPI_ISL_659922\|20 | ···················· | ···················· | ····················· | ···················· | ···················· | ···················· |
| S | 6357 hCoV-19/England/QEUH-B2F9C9/2020\|EPI_ISL_659923\|20 | ···················· | ···················· | ····················· | ···················· | ···················· | ···················· |
| S | 6358 hCoV-19/England/QEUH-B2D3CD/2020\|EPI_ISL_659924\|20 | ···················· | ···················· | ····················· | ···················· | ···················· | ···················· |
| S | 6359 hCoV-19/England/QEUH-B30330/2020\|EPI_ISL_659971\|20 | ···················· | ···················· | ····················· | ···················· | ···················· | ···················· |
| S | 6360 hCoV-19/England/QEUH-B2F5D4/2020\|EPI_ISL_659977\|20 | ···················· | ···················· | ····················· | ···················· | ···················· | ···················· |
| S | 6361 hCoV-19/England/QEUH-B2F8F9/2020\|EPI_ISL_659978\|20 | ···················· | ···················· | ····················· | ···················· | ···················· | ···················· |
| S | 6362 hCoV-19/England/QEUH-B2D56D/2020\|EPI_ISL_659979\|20 | ···················· | ···················· | ····················· | ···················· | ···················· | ···················· |
| S | 6363 hCoV-19/England/QEUH-B2CD57/2020\|EPI_ISL_659991\|20 | ···················· | ···················· | ····················· | ···················· | ···················· | ···················· |
| S | 6364 hCoV-19/England/QEUH-B2EB2E/2020\|EPI_ISL_659993\|20 | ···················· | ···················· | ····················· | ···················· | ···················· | ···················· |
| S | 6365 hCoV-19/England/QEUH-B2F99C/2020\|EPI_ISL_659994\|20 | ···················· | ···················· | ····················· | ···················· | ···················· | ···················· |
| S | 6366 hCoV-19/England/QEUH-B2CCC3/2020\|EPI_ISL_659996\|20 | ···················· | ···················· | ····················· | ···················· | ···················· | ···················· |
| S | 6367 hCoV-19/England/QEUH-B30A86/2020\|EPI_ISL_659997\|20 | ···················· | ···················· | ····················· | ···················· | ···················· | ···················· |
| S | 6368 hCoV-19/England/QEUH-B2D248/2020\|EPI_ISL_659998\|20 | ···················· | ···················· | ····················· | ···················· | ···················· | ···················· |
| S | 6369 hCoV-19/USA/WI-GMF-24235/2020\|EPI_ISL_660900\|2020- | ···················· | ···················· | ····················· | ···················· | ···················· | ···················· |
| S | 6370 hCoV-19/USA/WI-GMF-24330/2020\|EPI_ISL_660901\|2020- | ···················· | ···················· | ····················· | ···················· | ···················· | ···················· |
| S | 6371 hCoV-19/USA/IA-GMF-24556/2020\|EPI_ISL_660902\|2020- | ···················· | ···················· | ····················· | ···················· | ···················· | ···················· |
| S | 6372 hCoV-19/USA/WI-GMF-24579/2020\|EPI_ISL_660903\|2020- | ···················· | ···················· | ····················· | ···················· | ···················· | ···················· |
| S | 6373 hCoV-19/USA/WI-GMF-24597/2020\|EPI_ISL_660904\|2020- | ···················· | ···················· | ····················· | ···················· | ···················· | ···················· |
| S | 6374 hCoV-19/USA/WI-GMF-24599/2020\|EPI_ISL_660905\|2020- | ···················· | ···················· | ····················· | ···················· | ···················· | ···················· |
| S | 6375 hCoV-19/USA/WI-GMF-24611/2020\|EPI_ISL_660906\|2020- | ···················· | ···················· | ····················· | ···················· | ···················· | ···················· |
| S | 6376 hCoV-19/USA/WI-GMF-24693/2020\|EPI_ISL_660907\|2020- | ···················· | ···················· | ····················· | ···················· | ···················· | ···················· |
| S | 6377 hCoV-19/USA/WI-GMF-24736/2020\|EPI_ISL_660908\|2020- | ···················· | ···················· | ····················· | ···················· | ···················· | ···················· |
| S | 6378 hCoV-19/USA/WI-GMF-24746/2020\|EPI_ISL_660909\|2020- | ···················· | ···················· | ····················· | ···················· | ···················· | ···················· |
| S | 6379 hCoV-19/USA/WI-GMF-27162/2020\|EPI_ISL_660970\|2020- | ···················· | ···················· | ····················· | ···················· | ···················· | ···················· |
| S | 6380 hCoV-19/USA/WI-GMF-27181/2020\|EPI_ISL_660971\|2020- | ···················· | ···················· | ····················· | ···················· | ···················· | ···················· |
| S | 6381 hCoV-19/USA/WI-GMF-27189/2020\|EPI_ISL_660972\|2020- | ···················· | ···················· | ····················· | ···················· | ···················· | ···················· |
| S | 6382 hCoV-19/USA/WI-GMF-27202/2020\|EPI_ISL_660973\|2020- | ···················· | ···················· | ····················· | ···················· | ···················· | ···················· |
| S | 6383 hCoV-19/USA/WI-GMF-27242/2020\|EPI_ISL_660974\|2020- | ···················· | ···················· | ····················· | ···················· | ···················· | ···················· |
| S | 6384 hCoV-19/USA/WI-GMF-27309/2020\|EPI_ISL_660975\|2020- | ···················· | ···················· | ····················· | ···················· | ···················· | ···················· |
| S | 6385 hCoV-19/USA/WI-GMF-27343/2020\|EPI_ISL_660976\|2020- | ···················· | ···················· | ····················· | ···················· | ···················· | ···················· |
| S | 6386 hCoV-19/USA/WI-GMF-27474/2020\|EPI_ISL_660977\|2020- | ···················· | ···················· | ····················· | ···················· | ···················· | ···················· |
| S | 6387 hCoV-19/USA/WI-GMF-27490/2020\|EPI_ISL_660978\|2020- | ···················· | ···················· | ····················· | ···················· | ···················· | ···················· |
| S | 6388 hCoV-19/USA/WI-GMF-27504/2020\|EPI_ISL_660979\|2020- | ···················· | ···················· | ····················· | ···················· | ···················· | ···················· |
| S | 6389 hCoV-19/USA/WI-GMF-27517/2020\|EPI_ISL_660980\|2020- | ···················· | ···················· | ····················· | ···················· | ···················· | ···················· |
| S | 6390 hCoV-19/USA/WI-GMF-27569/2020\|EPI_ISL_660981\|2020- | ···················· | ···················· | ····················· | ···················· | ···················· | ···················· |
| S | 6391 hCoV-19/USA/WI-GMF-27617/2020\|EPI_ISL_660982\|2020- | ···················· | ···················· | ····················· | ···················· | ···················· | ···················· |
| S | 6392 hCoV-19/USA/WI-GMF-27637/2020\|EPI_ISL_660983\|2020- | ···················· | ·A·················· | ····················· | ···················· | ···················· | ···················· |
| S | 6393 hCoV-19/USA/WI-GMF-27653/2020\|EPI_ISL_660984\|2020- | ···················· | ···················· | ····················· | ···················· | ···················· | ···················· |
| S | 6394 hCoV-19/USA/WI-GMF-27681/2020\|EPI_ISL_660985\|2020- | ···················· | ···················· | ····················· | ···················· | ···················· | ···················· |
| S | 6395 hCoV-19/USA/WI-GMF-27686/2020\|EPI_ISL_660986\|2020- | ···················· | ···················· | ····················· | ···················· | ···················· | ···················· |
| S | 6396 hCoV-19/USA/IA-GMF-27699/2020\|EPI_ISL_660987\|2020- | ···················· | ···················· | ····················· | ···················· | ···················· | ···················· |
| S | 6397 hCoV-19/USA/MN-GMF-27759/2020\|EPI_ISL_660988\|2020- | ···················· | ·A·················· | ····················· | ···················· | ···················· | ···················· |
| S | 6398 hCoV-19/USA/WI-GMF-27773/2020\|EPI_ISL_660989\|2020- | ···················· | ···················· | ····················· | ···················· | ···················· | ···················· |
| S | 6399 hCoV-19/USA/MN-GMF-27790/2020\|EPI_ISL_660990\|2020- | ···················· | ···················· | ····················· | ···················· | ···················· | ···················· |
| S | 6400 hCoV-19/USA/WI-GMF-27794/2020\|EPI_ISL_660991\|2020- | ···················· | ···················· | ····················· | ···················· | ···················· | ···················· |
| S | 6402 hCoV-19/Scotland/QEUH-AE28B7/2020\|EPI_ISL_649651\|2 | ···················· | ···················· | ····················· | ···················· | ···················· | ···················· |
| S | 6403 hCoV-19/Scotland/QEUH-AE26AE/2020\|EPI_ISL_649652\|2 | ···················· | ···················· | ····················· | ···················· | ···················· | ···················· |
| S | 6404 hCoV-19/Scotland/QEUH-AE2750/2020\|EPI_ISL_649653\|2 | ···················· | ···················· | ····················· | ···················· | ···················· | ···················· |
| S | 6405 hCoV-19/Scotland/QEUH-AE2E79/2020\|EPI_ISL_649654\|2 | ···················· | ···················· | ····················· | ···················· | ···················· | ···················· |
| S | 6406 hCoV-19/Scotland/QEUH-AE1FB3/2020\|EPI_ISL_649655\|2 | ···················· | ···················· | ····················· | ···················· | ···················· | ···················· |
| S | 6407 hCoV-19/Scotland/QEUH-AE2125/2020\|EPI_ISL_649656\|2 | ···················· | ···················· | ····················· | ···················· | ···················· | ···················· |
| S | 6408 hCoV-19/Scotland/QEUH-AE1E2F/2020\|EPI_ISL_649657\|2 | ···················· | ···················· | ····················· | ···················· | ···················· | ···················· |
| S | 6409 hCoV-19/Scotland/QEUH-AE1F86/2020\|EPI_ISL_649658\|2 | ···················· | ···················· | ····················· | ···················· | ···················· | ···················· |
| S | 6410 hCoV-19/Scotland/QEUH-AE278D/2020\|EPI_ISL_649659\|2 | ···················· | ···················· | ····················· | ···················· | ···················· | ···················· |
| S | 6411 hCoV-19/Scotland/QEUH-AE49EF/2020\|EPI_ISL_649670\|2 | ···················· | ···················· | ····················· | ···················· | ···················· | ···················· |
| S | 6412 hCoV-19/Scotland/QEUH-AD101D/2020\|EPI_ISL_649671\|2 | ···················· | ···················· | ····················· | ···················· | ···················· | ···················· |
| S | 6413 hCoV-19/Scotland/QEUH-AD542C/2020\|EPI_ISL_649672\|2 | ···················· | ···················· | ····················· | ···················· | ···················· | ···················· |
| S | 6414 hCoV-19/Scotland/QEUH-AD0F7B/2020\|EPI_ISL_649673\|2 | ···················· | ···················· | ····················· | ···················· | ···················· | ···················· |
| S | 6415 hCoV-19/Scotland/QEUH-AD5B27/2020\|EPI_ISL_649675\|2 | ···················· | ···················· | ····················· | ···················· | ···················· | ···················· |
| S | 6416 hCoV-19/Scotland/QEUH-AD16A2/2020\|EPI_ISL_649676\|2 | ···················· | ···················· | ····················· | ···················· | ···················· | ···················· |
| S | 6417 hCoV-19/Scotland/QEUH-AD1903/2020\|EPI_ISL_649677\|2 | ···················· | ···················· | ····················· | ···················· | ···················· | ···················· |
| S | 6418 hCoV-19/Scotland/QEUH-AD53F2/2020\|EPI_ISL_649678\|2 | ···················· | ···················· | ····················· | ···················· | ···················· | ···················· |
| S | 6419 hCoV-19/Scotland/QEUH-AD5477/2020\|EPI_ISL_649679\|2 | ···················· | ···················· | ····················· | ···················· | ···················· | ···················· |
| S | 6420 hCoV-19/Scotland/QEUH-AD5714/2020\|EPI_ISL_649680\|2 | ···················· | ···················· | ····················· | ···················· | ···················· | ···················· |

| | | | | | | | | |
|---|---|---|---|---|---|---|---|---|
| S | 6421 | hCoV-19/Scotland/QEUH-AD5BCD/2020\|EPI_ISL_649681\|2 | ···················· | ···················· | ···················· | ···················· | ···················· | ···················· |
| S | 6422 | hCoV-19/Scotland/QEUH-AD435D/2020\|EPI_ISL_649682\|2 | ···················· | ···················· | ···················· | ···················· | ···················· | ···················· |
| S | 6423 | hCoV-19/Scotland/QEUH-AD1684/2020\|EPI_ISL_649683\|2 | ···················· | ···················· | ···················· | ···················· | ···················· | ···················· |
| S | 6424 | hCoV-19/Scotland/QEUH-AD0FD5/2020\|EPI_ISL_649684\|2 | ···················· | ···················· | ···················· | ···················· | ···················· | ···················· |
| S | 6425 | hCoV-19/Scotland/QEUH-AD1499/2020\|EPI_ISL_649685\|2 | ···················· | ···················· | ···················· | ···················· | ···················· | ···················· |
| S | 6426 | hCoV-19/Scotland/QEUH-AD14A8/2020\|EPI_ISL_649686\|2 | ···················· | ···················· | ···················· | ···················· | ···················· | ···················· |
| S | 6427 | hCoV-19/Scotland/QEUH-AD1657/2020\|EPI_ISL_649687\|2 | ···················· | ···················· | ···················· | ···················· | ···················· | ···················· |
| S | 6428 | hCoV-19/Scotland/QEUH-AD57D8/2020\|EPI_ISL_649688\|2 | ···················· | ·A·················· | ···················· | ···················· | ···················· | ···················· |
| S | 6429 | hCoV-19/Scotland/QEUH-AD56CC/2020\|EPI_ISL_649689\|2 | ···················· | ···················· | ···················· | ···················· | ···················· | ···················· |
| S | 6430 | hCoV-19/Scotland/EDB8565/2020\|EPI_ISL_651961\|2020- | ···················· | ···················· | ···················· | ···················· | ···················· | ···················· |
| S | 6435 | hCoV-19/Scotland/EDB8585/2020\|EPI_ISL_651966\|2020- | ···················· | ···················· | ···················· | ···················· | ···················· | ···················· |
| S | 6436 | hCoV-19/Scotland/EDB8586/2020\|EPI_ISL_651967\|2020- | ···················· | ···················· | ···················· | ···················· | ···················· | ···················· |
| S | 6438 | hCoV-19/Scotland/EDB8588/2020\|EPI_ISL_651969\|2020- | ···················· | ···················· | ···················· | ···················· | ·····T·············· | ···················· |
| S | 6442 | hCoV-19/Scotland/EDB8600/2020\|EPI_ISL_651975\|2020- | ···················· | ···················· | ···················· | ···················· | ···················· | ···················· |
| S | 6443 | hCoV-19/Scotland/EDB8614/2020\|EPI_ISL_651980\|2020- | ···················· | ···················· | ···················· | ···················· | ···················· | ···················· |
| S | 6444 | hCoV-19/Scotland/EDB8615/2020\|EPI_ISL_651981\|2020- | ···················· | ···················· | ···················· | ···················· | ···················· | ···················· |
| S | 6445 | hCoV-19/Scotland/EDB8626/2020\|EPI_ISL_651982\|2020- | ···················· | ···················· | ···················· | ···················· | ···················· | ···················· |
| S | 6446 | hCoV-19/Scotland/EDB8627/2020\|EPI_ISL_651983\|2020- | ···················· | ···················· | ···················· | ···················· | ···················· | ···················· |
| S | 6447 | hCoV-19/Scotland/EDB8630/2020\|EPI_ISL_651984\|2020- | ···················· | ···················· | ···················· | ···················· | ···················· | ···················· |
| S | 6448 | hCoV-19/Scotland/EDB8640/2020\|EPI_ISL_651985\|2020- | ···················· | ···················· | ···················· | ···················· | ···················· | ···················· |
| S | 6449 | hCoV-19/Scotland/EDB8641/2020\|EPI_ISL_651986\|2020- | ···················· | ···················· | ···················· | ···················· | ···················· | ···················· |
| S | 6450 | hCoV-19/Scotland/EDB8678/2020\|EPI_ISL_651987\|2020- | ···················· | ···················· | ···················· | ···················· | ···················· | ···················· |
| S | 6451 | hCoV-19/Scotland/EDB8679/2020\|EPI_ISL_651988\|2020- | ···················· | ···················· | ···················· | ···················· | ···················· | ···················· |
| S | 6452 | hCoV-19/Scotland/EDB8745/2020\|EPI_ISL_651989\|2020- | ···················· | ···················· | ···················· | ···················· | ···················· | ···················· |
| S | 6453 | hCoV-19/Scotland/EDB8748/2020\|EPI_ISL_651990\|2020- | ···················· | ···················· | ···················· | ···················· | ···················· | ···················· |
| S | 6454 | hCoV-19/Scotland/EDB8754/2020\|EPI_ISL_651991\|2020- | ···················· | ···················· | ···················· | ···················· | ···················· | ···················· |
| S | 6455 | hCoV-19/Scotland/EDB8756/2020\|EPI_ISL_651992\|2020- | ···················· | ···················· | ···················· | ···················· | ···················· | ···················· |
| S | 6456 | hCoV-19/Scotland/EDB8769/2020\|EPI_ISL_651993\|2020- | ···················· | ···················· | ···················· | ···················· | ···················· | ···················· |
| S | 6458 | hCoV-19/Scotland/EDB8776/2020\|EPI_ISL_651995\|2020- | ···················· | ···················· | ···················· | ···················· | ···················· | ···················· |
| S | 6459 | hCoV-19/Scotland/EDB8783/2020\|EPI_ISL_651996\|2020- | ···················· | ···················· | ···················· | ···················· | ···················· | ···················· |
| S | 6460 | hCoV-19/Scotland/EDB8787/2020\|EPI_ISL_651997\|2020- | ···················· | ···················· | ···················· | ···················· | ···················· | ···················· |
| S | 6461 | hCoV-19/Scotland/EDB8792/2020\|EPI_ISL_651998\|2020- | ···················· | ···················· | ···················· | ···················· | ···················· | ···················· |
| S | 6462 | hCoV-19/Scotland/EDB8794/2020\|EPI_ISL_651999\|2020- | ···················· | ···················· | ···················· | ···················· | ···················· | ···················· |
| S | 6463 | hCoV-19/England/QEUH-B433EF/2020\|EPI_ISL_657300\|20 | ···················· | ···················· | ···················· | ···················· | ···················· | ···················· |
| S | 6464 | hCoV-19/England/QEUH-B433D0/2020\|EPI_ISL_657301\|20 | ···················· | ···················· | ···················· | ···················· | ···················· | ···················· |
| S | 6465 | hCoV-19/England/QEUH-B437F2/2020\|EPI_ISL_657302\|20 | ···················· | ···················· | ···················· | ···················· | ···················· | ···················· |
| S | 6466 | hCoV-19/England/QEUH-B43613/2020\|EPI_ISL_657303\|20 | ···················· | ···················· | ···················· | ···················· | ···················· | ···················· |
| S | 6467 | hCoV-19/England/QEUH-B3FF3D/2020\|EPI_ISL_657304\|20 | ···················· | ···················· | ···················· | ···················· | ···················· | ···················· |
| S | 6468 | hCoV-19/England/QEUH-B421AA/2020\|EPI_ISL_657305\|20 | ···················· | ···················· | ···················· | ···················· | ···················· | ···················· |
| S | 6469 | hCoV-19/England/QEUH-B4365F/2020\|EPI_ISL_657306\|20 | ···················· | ···················· | ···················· | ···················· | ···················· | ···················· |
| S | 6470 | hCoV-19/England/QEUH-B4376B/2020\|EPI_ISL_657307\|20 | ···················· | ···················· | ···················· | ···················· | ···················· | ···················· |
| S | 6471 | hCoV-19/England/QEUH-B408A7/2020\|EPI_ISL_657308\|20 | ···················· | ···················· | ···················· | ···················· | ···················· | ···················· |
| S | 6472 | hCoV-19/England/QEUH-B427E4/2020\|EPI_ISL_657309\|20 | ···················· | ···················· | ···················· | ···················· | ···················· | ···················· |
| S | 6473 | hCoV-19/England/QEUH-B423D1/2020\|EPI_ISL_657311\|20 | ···················· | ···················· | ···················· | ···················· | ···················· | ···················· |
| S | 6474 | hCoV-19/England/QEUH-B4369B/2020\|EPI_ISL_657312\|20 | ···················· | ···················· | ···················· | ···················· | ···················· | ···················· |
| S | 6475 | hCoV-19/England/QEUH-B402D6/2020\|EPI_ISL_657313\|20 | ···················· | ···················· | ···················· | ···················· | ···················· | ···················· |
| S | 6476 | hCoV-19/England/QEUH-B420E9/2020\|EPI_ISL_657318\|20 | ···················· | ···················· | ···················· | ···················· | ·····T·············· | ···················· |
| S | 6477 | hCoV-19/England/QEUH-B43947/2020\|EPI_ISL_657319\|20 | ···················· | ···················· | ···················· | ···················· | ···················· | ···················· |
| S | 6478 | hCoV-19/England/QEUH-B422D4/2020\|EPI_ISL_657320\|20 | ···················· | ···················· | ···················· | ···················· | ···················· | ···················· |
| S | 6479 | hCoV-19/England/QEUH-B438C2/2020\|EPI_ISL_657321\|20 | ···················· | ···················· | ···················· | ···················· | ···················· | ···················· |
| S | 6480 | hCoV-19/England/QEUH-B40133/2020\|EPI_ISL_657322\|20 | ···················· | ···················· | ···················· | ···················· | ···················· | ···················· |
| S | 6481 | hCoV-19/England/QEUH-B43482/2020\|EPI_ISL_657323\|20 | ···················· | ···················· | ···················· | ···················· | ·····T·············· | ···················· |
| S | 6482 | hCoV-19/England/QEUH-B40449/2020\|EPI_ISL_657324\|20 | ···················· | ···················· | ···················· | ···················· | ···················· | ···················· |
| S | 6483 | hCoV-19/England/QEUH-B408D4/2020\|EPI_ISL_657325\|20 | ···················· | ···················· | ···················· | ···················· | ···················· | ···················· |
| S | 6484 | hCoV-19/England/QEUH-B427F3/2020\|EPI_ISL_657326\|20 | ···················· | ···················· | ···················· | ···················· | ···················· | ···················· |
| S | 6485 | hCoV-19/England/QEUH-B40977/2020\|EPI_ISL_657327\|20 | ···················· | ···················· | ···················· | ···················· | ···················· | ···················· |
| S | 6486 | hCoV-19/England/QEUH-B42526/2020\|EPI_ISL_657328\|20 | ···················· | ···················· | ···················· | ···················· | ···················· | ···················· |
| S | 6487 | hCoV-19/England/QEUH-B3F850/2020\|EPI_ISL_658600\|20 | ···················· | ···················· | ···················· | ···················· | ···················· | ···················· |
| S | 6488 | hCoV-19/England/QEUH-B3FD60/2020\|EPI_ISL_658601\|20 | ···················· | ···················· | ···················· | ···················· | ···················· | ··A················· |
| S | 6489 | hCoV-19/England/QEUH-B3F999/2020\|EPI_ISL_658602\|20 | ···················· | ···················· | ···················· | ···················· | ···················· | ···················· |
| S | 6490 | hCoV-19/England/QEUH-B3F692/2020\|EPI_ISL_658603\|20 | ···················· | ·A·················· | ···················· | ···················· | ···················· | ···················· |
| S | 6491 | hCoV-19/England/QEUH-B41721/2020\|EPI_ISL_658604\|20 | ···················· | ···················· | ···················· | ···················· | ···················· | ···················· |
| S | 6492 | hCoV-19/England/QEUH-B41800/2020\|EPI_ISL_658605\|20 | ···················· | ···················· | ···················· | ···················· | ···················· | ···················· |
| S | 6493 | hCoV-19/England/QEUH-B3F3AA/2020\|EPI_ISL_658606\|20 | ···················· | ···················· | ···················· | ···················· | ···················· | ···················· |
| S | 6494 | hCoV-19/England/QEUH-B3F93F/2020\|EPI_ISL_658607\|20 | ···················· | ···················· | ···················· | ···················· | ···················· | ···················· |
| S | 6495 | hCoV-19/England/QEUH-B41581/2020\|EPI_ISL_658608\|20 | ···················· | ···················· | ···················· | ···················· | ···················· | ···················· |
| S | 6496 | hCoV-19/England/QEUH-B417C7/2020\|EPI_ISL_658609\|20 | ···················· | ···················· | ···················· | ···················· | ···················· | ···················· |
| S | 6497 | hCoV-19/England/QEUH-B3F6ED/2020\|EPI_ISL_658610\|20 | ···················· | ···················· | ···················· | ···················· | ···················· | ···················· |
| S | 6498 | hCoV-19/England/QEUH-B3F60B/2020\|EPI_ISL_658611\|20 | ···················· | ···················· | ···················· | ···················· | ···················· | ···················· |
| S | 6499 | hCoV-19/England/QEUH-B3F674/2020\|EPI_ISL_658612\|20 | ···················· | ···················· | ···················· | ···················· | ···················· | ···················· |
| S | 6500 | hCoV-19/England/QEUH-B412E4/2020\|EPI_ISL_658613\|20 | ···················· | ···················· | ···················· | ···················· | ···················· | ···················· |
| S | 6501 | hCoV-19/England/QEUH-B418A6/2020\|EPI_ISL_658614\|20 | ···················· | ···················· | ···················· | ···················· | ···················· | ···················· |
| S | 6502 | hCoV-19/England/QEUH-B3F401/2020\|EPI_ISL_658615\|20 | ···················· | ···················· | ···················· | ···················· | ···················· | ···················· |
| S | 6503 | hCoV-19/England/QEUH-B3FDAC/2020\|EPI_ISL_658616\|20 | ···················· | ···················· | ···················· | ···················· | ···················· | ···················· |
| S | 6504 | hCoV-19/England/QEUH-B3F647/2020\|EPI_ISL_658617\|20 | ···················· | ···················· | ···················· | ···················· | ···················· | ···················· |
| S | 6505 | hCoV-19/England/QEUH-B41518/2020\|EPI_ISL_658618\|20 | ···················· | ···················· | ···················· | ···················· | ···················· | ···················· |
| S | 6506 | hCoV-19/England/QEUH-B41642/2020\|EPI_ISL_658619\|20 | ···················· | ···················· | ···················· | ···················· | ···················· | ···················· |
| S | 6507 | hCoV-19/England/QEUH-B4181F/2020\|EPI_ISL_658620\|20 | ···················· | ···················· | ···················· | ···················· | ···················· | ···················· |
| S | 6508 | hCoV-19/England/QEUH-B3FB1B/2020\|EPI_ISL_658621\|20 | ···················· | ···················· | ···················· | ···················· | ···················· | ···················· |
| S | 6509 | hCoV-19/England/QEUH-B41C7D/2020\|EPI_ISL_658622\|20 | ···················· | ···················· | ···················· | ···················· | ···················· | ···················· |

| | | | | | | | |
|---|---|---|---|---|---|---|---|
| S | 6510 hCoV-19/England/QEUH-B3FA4B/2020\|EPI_ISL_658623\|20 | ···················· | ···················· | ····················· | ···················· | ···················· | ···················· |
| S | 6511 hCoV-19/England/QEUH-B4183D/2020\|EPI_ISL_658624\|20 | ···················· | ···················· | ····················· | ···················· | ···················· | ···················· |
| S | 6512 hCoV-19/England/QEUH-B3FC45/2020\|EPI_ISL_658625\|20 | ···················· | ···················· | ····················· | ···················· | ···················· | ···················· |
| S | 6513 hCoV-19/England/QEUH-B419C1/2020\|EPI_ISL_658626\|20 | ···················· | ···················· | ····················· | ···················· | ···················· | ···················· |
| S | 6514 hCoV-19/England/QEUH-B4179A/2020\|EPI_ISL_658627\|20 | ···················· | ···················· | ····················· | ···················· | ···················· | ···················· |
| S | 6515 hCoV-19/England/QEUH-B3F629/2020\|EPI_ISL_658628\|20 | ···················· | ···················· | ····················· | ···················· | ···················· | ···················· |
| S | 6516 hCoV-19/England/QEUH-B3F7DB/2020\|EPI_ISL_658629\|20 | ···················· | ···················· | ····················· | ···················· | ···················· | ···················· |
| S | 6517 hCoV-19/England/QEUH-B3F5C2/2020\|EPI_ISL_658630\|20 | ···················· | ···················· | ····················· | ···················· | ···················· | ···················· |
| S | 6518 hCoV-19/England/QEUH-B4127B/2020\|EPI_ISL_658631\|20 | ···················· | ···················· | ····················· | ···················· | ···················· | ···················· |
| S | 6519 hCoV-19/England/QEUH-B41A55/2020\|EPI_ISL_658632\|20 | ···················· | ···················· | ····················· | ···················· | ···················· | ···················· |
| S | 6520 hCoV-19/England/QEUH-B41457/2020\|EPI_ISL_658633\|20 | ···················· | ···················· | ····················· | ···················· | ···················· | ···················· |
| S | 6521 hCoV-19/England/QEUH-B41958/2020\|EPI_ISL_658634\|20 | ···················· | ···················· | ····················· | ···················· | ···················· | ···················· |
| S | 6522 hCoV-19/England/QEUH-B419D0/2020\|EPI_ISL_658635\|20 | ···················· | ···················· | ····················· | ···················· | ···················· | ···················· |
| S | 6523 hCoV-19/England/QEUH-B41035/2020\|EPI_ISL_658636\|20 | ···················· | ···················· | ····················· | ···················· | ···················· | ···················· |
| S | 6524 hCoV-19/England/QEUH-B4118D/2020\|EPI_ISL_658637\|20 | ···················· | ···················· | ····················· | ···················· | ···················· | ···················· |
| S | 6525 hCoV-19/England/QEUH-B4177C/2020\|EPI_ISL_658638\|20 | ···················· | ···················· | ····················· | ···················· | ···················· | ···················· |
| S | 6526 hCoV-19/England/QEUH-B3F744/2020\|EPI_ISL_658641\|20 | ···················· | ···················· | ····················· | ···················· | ···················· | ···················· |
| S | 6527 hCoV-19/England/QEUH-B3F270/2020\|EPI_ISL_658643\|20 | ···················· | ···················· | ····················· | ···················· | ···················· | ···················· |
| S | 6528 hCoV-19/England/QEUH-B3F489/2020\|EPI_ISL_658644\|20 | ···················· | ···················· | ····················· | ···················· | ···················· | ···················· |
| S | 6529 hCoV-19/England/QEUH-B3F683/2020\|EPI_ISL_658649\|20 | ···················· | ···················· | ····················· | ···················· | ···················· | ···················· |
| S | 6530 hCoV-19/England/QEUH-B41AFB/2020\|EPI_ISL_658650\|20 | ···················· | ···················· | ····················· | ···················· | ···················· | ···················· |
| S | 6531 hCoV-19/England/QEUH-B3F7F9/2020\|EPI_ISL_658651\|20 | ···················· | ···················· | ····················· | ···················· | ···················· | ···················· |
| S | 6532 hCoV-19/England/QEUH-B3F753/2020\|EPI_ISL_658652\|20 | ···················· | ···················· | ····················· | ···················· | ···················· | ···················· |
| S | 6533 hCoV-19/England/QEUH-B3F559/2020\|EPI_ISL_658653\|20 | ···················· | ·A·················· | ····················· | ···················· | ···················· | ···················· |
| S | 6534 hCoV-19/England/QEUH-B3FA1E/2020\|EPI_ISL_658654\|20 | ···················· | ···················· | ····················· | ···················· | ···················· | ···················· |
| S | 6535 hCoV-19/England/QEUH-B40F75/2020\|EPI_ISL_658656\|20 | ···················· | ···················· | ····················· | ···················· | ···················· | ···················· |
| S | 6536 hCoV-19/England/QEUH-B41606/2020\|EPI_ISL_658657\|20 | ···················· | ···················· | ····················· | ···················· | ···················· | ···················· |
| S | 6537 hCoV-19/England/QEUH-B415FA/2020\|EPI_ISL_658658\|20 | ···················· | ···················· | ····················· | ···················· | ···················· | ···················· |
| S | 6538 hCoV-19/England/QEUH-B411E7/2020\|EPI_ISL_658659\|20 | ···················· | ···················· | ····················· | ···················· | ···················· | ···················· |
| S | 6539 hCoV-19/England/CAMC-B37FBD/2020\|EPI_ISL_659000\|20 | ···················· | ···················· | ····················· | ···················· | ···················· | ···················· |
| S | 6540 hCoV-19/England/MILK-B3958C/2020\|EPI_ISL_659001\|20 | ···················· | ···················· | ····················· | ···················· | ···················· | ··A················· |
| S | 6541 hCoV-19/England/MILK-B385BA/2020\|EPI_ISL_659002\|20 | ···················· | ···················· | ····················· | ···················· | ···················· | ···················· |
| S | 6542 hCoV-19/England/MILK-B386C6/2020\|EPI_ISL_659004\|20 | ···················· | ···················· | ····················· | ···················· | ···················· | ···················· |
| S | 6543 hCoV-19/England/CAMC-B37D59/2020\|EPI_ISL_659005\|20 | ···················· | ···················· | ····················· | ···················· | ···················· | ···················· |
| S | 6544 hCoV-19/England/MILK-B383FC/2020\|EPI_ISL_659006\|20 | ···················· | ···················· | ····················· | ···················· | ···················· | ···················· |
| S | 6545 hCoV-19/England/CAMC-B380F5/2020\|EPI_ISL_659007\|20 | ···················· | ···················· | ····················· | ···················· | ···················· | ···················· |
| S | 6546 hCoV-19/England/MILK-B394F8/2020\|EPI_ISL_659008\|20 | ···················· | ···················· | ····················· | ···················· | ···················· | ···················· |
| S | 6547 hCoV-19/England/MILK-B3A57C/2020\|EPI_ISL_659009\|20 | ···················· | ···················· | ····················· | ···················· | ···················· | ···················· |
| S | 6548 hCoV-19/England/QEUH-B2FCFD/2020\|EPI_ISL_659925\|20 | ···················· | ···················· | ····················· | ···················· | ···················· | ···················· |
| S | 6549 hCoV-19/England/QEUH-B2F6D1/2020\|EPI_ISL_659926\|20 | ···················· | ···················· | ····················· | ···················· | ···················· | ···················· |
| S | 6550 hCoV-19/England/QEUH-B2E836/2020\|EPI_ISL_659927\|20 | ···················· | ···················· | ····················· | ···················· | ···················· | ···················· |
| S | 6551 hCoV-19/England/QEUH-B2DF6F/2020\|EPI_ISL_659928\|20 | ···················· | ···················· | ····················· | ···················· | ···················· | ···················· |
| S | 6552 hCoV-19/England/QEUH-B2F8AE/2020\|EPI_ISL_659929\|20 | ···················· | ···················· | ····················· | ···················· | ···················· | ···················· |
| S | 6553 hCoV-19/England/QEUH-B2EDDD/2020\|EPI_ISL_659930\|20 | ···················· | ···················· | ····················· | ···················· | ···················· | ···················· |
| S | 6554 hCoV-19/England/QEUH-B2D679/2020\|EPI_ISL_659931\|20 | ···················· | ···················· | ····················· | ···················· | ···················· | ···················· |
| S | 6555 hCoV-19/England/QEUH-B2D8BF/2020\|EPI_ISL_659932\|20 | ···················· | ···················· | ····················· | ···················· | ···················· | ···················· |
| S | 6556 hCoV-19/England/QEUH-B2E809/2020\|EPI_ISL_659933\|20 | ···················· | ·A·················· | ····················· | ···················· | ···················· | ···················· |
| S | 6557 hCoV-19/England/QEUH-B303B8/2020\|EPI_ISL_659934\|20 | ···················· | ···················· | ····················· | ···················· | ···················· | ···················· |
| S | 6558 hCoV-19/England/QEUH-B2D381/2020\|EPI_ISL_659935\|20 | ···················· | ···················· | ····················· | ···················· | ···················· | ···················· |
| S | 6559 hCoV-19/England/QEUH-B2EEBC/2020\|EPI_ISL_659936\|20 | ···················· | ···················· | ····················· | ···················· | ···················· | ···············A···· |
| S | 6560 hCoV-19/England/QEUH-B2D837/2020\|EPI_ISL_659937\|20 | ···················· | ···················· | ····················· | ···················· | ···················· | ···················· |
| S | 6561 hCoV-19/England/QEUH-B2EEAD/2020\|EPI_ISL_659938\|20 | ···················· | ···················· | ····················· | ···················· | ···················· | ···················· |
| S | 6562 hCoV-19/England/QEUH-B2D336/2020\|EPI_ISL_659939\|20 | ···················· | ···················· | ····················· | ···················· | ···················· | ···················· |
| S | 6563 hCoV-19/England/QEUH-B2F8DB/2020\|EPI_ISL_659940\|20 | ···················· | ···················· | ····················· | ···················· | ···················· | ···················· |
| S | 6564 hCoV-19/England/QEUH-B2F70B/2020\|EPI_ISL_659941\|20 | ···················· | ···················· | ····················· | ···················· | ···················· | ···················· |
| S | 6565 hCoV-19/England/QEUH-B2F5F2/2020\|EPI_ISL_659942\|20 | ···················· | ···················· | ····················· | ···················· | ···················· | ···················· |
| S | 6566 hCoV-19/England/QEUH-B30BED/2020\|EPI_ISL_659943\|20 | ···················· | ···················· | ····················· | ···················· | ···················· | ···················· |
| S | 6567 hCoV-19/England/QEUH-B2DEF9/2020\|EPI_ISL_659944\|20 | ···················· | ···················· | ····················· | ···················· | ···················· | ···················· |
| S | 6568 hCoV-19/England/QEUH-B30AC2/2020\|EPI_ISL_659945\|20 | ···················· | ···················· | ····················· | ···················· | ···················· | ···················· |
| S | 6569 hCoV-19/England/QEUH-B30AB3/2020\|EPI_ISL_659946\|20 | ···················· | ···················· | ····················· | ···················· | ···················· | ···················· |
| S | 6570 hCoV-19/England/QEUH-B2EAF4/2020\|EPI_ISL_659947\|20 | ···················· | ···················· | ····················· | ···················· | ···················· | ···················· |
| S | 6571 hCoV-19/England/QEUH-B3034F/2020\|EPI_ISL_659949\|20 | ···················· | ···················· | ····················· | ···················· | ···················· | ···················· |
| S | 6572 hCoV-19/England/QEUH-B2EE8F/2020\|EPI_ISL_659950\|20 | ···················· | ···················· | ····················· | ···················· | ···················· | ···················· |
| S | 6573 hCoV-19/England/QEUH-B2CE90/2020\|EPI_ISL_659951\|20 | ···················· | ···················· | ····················· | ···················· | ···················· | ···················· |
| S | 6574 hCoV-19/England/QEUH-B2D934/2020\|EPI_ISL_659953\|20 | ···················· | ···················· | ····················· | ···················· | ···················· | ···················· |
| S | 6575 hCoV-19/England/QEUH-B2D4BB/2020\|EPI_ISL_659955\|20 | ···················· | ···················· | ····················· | ···················· | ···················· | ···················· |
| S | 6576 hCoV-19/England/QEUH-B2DFAB/2020\|EPI_ISL_659956\|20 | ···················· | ···················· | ····················· | ···················· | ···················· | ···················· |
| S | 6577 hCoV-19/England/QEUH-B2EB79/2020\|EPI_ISL_659957\|20 | ···················· | ···················· | ····················· | ···················· | ···················· | ···················· |
| S | 6578 hCoV-19/England/QEUH-B2D828/2020\|EPI_ISL_659958\|20 | ···················· | ·········T·········· | ····················· | ···················· | ···················· | ···················· |
| S | 6579 hCoV-19/England/QEUH-B2D8DD/2020\|EPI_ISL_659959\|20 | ···················· | ···················· | ····················· | ···················· | ···················· | ···················· |
| S | 6580 hCoV-19/England/QEUH-B2D64C/2020\|EPI_ISL_659961\|20 | ···················· | ···················· | ····················· | ···················· | ···················· | ···················· |
| S | 6581 hCoV-19/England/QEUH-B2DEDB/2020\|EPI_ISL_659962\|20 | ···················· | ···················· | ····················· | ···················· | ···················· | ···················· |
| S | 6582 hCoV-19/England/QEUH-B30AFF/2020\|EPI_ISL_659963\|20 | ···················· | ···················· | ····················· | ···················· | ···················· | ···················· |
| S | 6583 hCoV-19/England/QEUH-B30479/2020\|EPI_ISL_659964\|20 | ···················· | ···················· | ····················· | ···················· | ···················· | ···················· |
| S | 6584 hCoV-19/England/QEUH-B2D952/2020\|EPI_ISL_659965\|20 | ···················· | ···················· | ····················· | ···················· | ···················· | ···················· |
| S | 6585 hCoV-19/England/QEUH-B2EB88/2020\|EPI_ISL_659966\|20 | ···················· | ···················· | ····················· | ···················· | ···················· | ···················· |
| S | 6586 hCoV-19/England/QEUH-B2EB5B/2020\|EPI_ISL_659967\|20 | ···················· | ···················· | ····················· | ···················· | ···················· | ···················· |
| S | 6587 hCoV-19/England/QEUH-B2EE16/2020\|EPI_ISL_659972\|20 | ···················· | ···················· | ····················· | ···················· | ···················· | ···················· |
| S | 6588 hCoV-19/England/QEUH-B2F729/2020\|EPI_ISL_659975\|20 | ···················· | ···················· | ····················· | ···················· | ···················· | ···················· |
| S | 6589 hCoV-19/England/QEUH-B2D58B/2020\|EPI_ISL_659980\|20 | ···················· | ···················· | ····················· | ···················· | ···················· | ···················· |

```
S  6590 hCoV-19/England/QEUH-B2D688/2020|EPI_ISL_659981|20   ····················  ····················  ····················  ····················  ····················  ····················
S  6591 hCoV-19/England/QEUH-B2E890/2020|EPI_ISL_659982|20   ····················  ····················  ····················  ····················  ····················  ····················
S  6592 hCoV-19/England/QEUH-B2EC67/2020|EPI_ISL_659983|20   ····················  ····················  ····················  ····················  ····················  ····················
S  6593 hCoV-19/England/QEUH-B30312/2020|EPI_ISL_659984|20   ····················  ····················  ····················  ····················  ····················  ····················
S  6594 hCoV-19/England/QEUH-B2CE81/2020|EPI_ISL_659986|20   ····················  ·A··················  ····················  ····················  ····················  ····················
S  6595 hCoV-19/England/QEUH-B2D3FA/2020|EPI_ISL_659988|20   ····················  ····················  ····················  ····················  ····················  ····················
S  6596 hCoV-19/England/QEUH-B2D309/2020|EPI_ISL_659989|20   ····················  ····················  ····················  ····················  ····················  ····················
S  6597 hCoV-19/USA/WI-GMF-24758/2020|EPI_ISL_660910|2020-   ····················  ····················  ····················  ····················  ····················  ····················
S  6598 hCoV-19/USA/WI-GMF-24763/2020|EPI_ISL_660911|2020-   ····················  ····················  ····················  ····················  ····················  ····················
S  6599 hCoV-19/USA/WI-GMF-24773/2020|EPI_ISL_660912|2020-   ····················  ····················  ····················  ····················  ····················  ····················
S  6600 hCoV-19/USA/WI-GMF-24788/2020|EPI_ISL_660913|2020-   ····················  ····················  ····················  ····················  ····················  ···············A····
S  6601 hCoV-19/USA/WI-GMF-24835/2020|EPI_ISL_660914|2020-   ····················  ····················  ····················  ····················  ····················  ····················
S  6602 hCoV-19/USA/WI-GMF-24867/2020|EPI_ISL_660915|2020-   ····················  ····················  ····················  ····················  ····················  ····················
S  6603 hCoV-19/USA/WI-GMF-25145/2020|EPI_ISL_660916|2020-   ····················  ····················  ····················  ····················  ····················  ····················
S  6604 hCoV-19/USA/IA-GMF-25157/2020|EPI_ISL_660917|2020-   ····················  ····················  ····················  ····················  ····················  ····················
S  6605 hCoV-19/USA/WI-GMF-25216/2020|EPI_ISL_660918|2020-   ····················  ····················  ····················  ····················  ····················  ····················
S  6606 hCoV-19/USA/WI-GMF-25226/2020|EPI_ISL_660919|2020-   ····················  ····················  ····················  ····················  ····················  ····················
S  6607 hCoV-19/USA/WI-GMF-25252/2020|EPI_ISL_660920|2020-   ····················  ····················  ····················  ····················  ····················  ····················
S  6608 hCoV-19/USA/WI-GMF-25260/2020|EPI_ISL_660921|2020-   ····················  ····················  ····················  ····················  ····················  ····················
S  6609 hCoV-19/USA/WI-GMF-25337/2020|EPI_ISL_660922|2020-   ····················  ····················  ····················  ····················  ····················  ····················
S  6610 hCoV-19/USA/WI-GMF-25355/2020|EPI_ISL_660923|2020-   ····················  ····················  ····················  ····················  ····················  ····················
S  6611 hCoV-19/USA/WI-GMF-25375/2020|EPI_ISL_660924|2020-   ····················  ····················  ····················  ····················  ····················  ····················
S  6612 hCoV-19/USA/WI-GMF-25385/2020|EPI_ISL_660925|2020-   ····················  ····················  ····················  ····················  ····················  ····················
S  6613 hCoV-19/USA/WI-GMF-25441/2020|EPI_ISL_660926|2020-   ····················  ····················  ····················  ····················  ····················  ····················
S  6614 hCoV-19/USA/WI-GMF-25446/2020|EPI_ISL_660927|2020-   ····················  ····················  ····················  ····················  ····················  ····················
S  6615 hCoV-19/USA/WI-GMF-25463/2020|EPI_ISL_660928|2020-   ····················  ····················  ····················  ····················  ····················  ····················
S  6616 hCoV-19/USA/WI-GMF-25469/2020|EPI_ISL_660929|2020-   ····················  ····················  ····················  ····················  ····················  ····················
S  6617 hCoV-19/USA/WI-GMF-25487/2020|EPI_ISL_660930|2020-   ····················  ····················  ····················  ····················  ····················  ····················
S  6618 hCoV-19/USA/WI-GMF-25500/2020|EPI_ISL_660931|2020-   ····················  ····················  ····················  ····················  ····················  ····················
S  6619 hCoV-19/USA/WI-GMF-25551/2020|EPI_ISL_660932|2020-   ····················  ····················  ····················  ····················  ····················  ····················
S  6620 hCoV-19/USA/WI-GMF-25643/2020|EPI_ISL_660933|2020-   ····················  ····················  ····················  ····················  ····················  ····················
S  6621 hCoV-19/USA/WI-GMF-25677/2020|EPI_ISL_660934|2020-   ····················  ····················  ····················  ····················  ····················  ····················
S  6622 hCoV-19/USA/WI-GMF-25854/2020|EPI_ISL_660935|2020-   ····················  ····················  ····················  ····················  ····················  ····················
S  6623 hCoV-19/USA/WI-GMF-25857/2020|EPI_ISL_660936|2020-   ····················  ····················  ····················  ····················  ····················  ····················
S  6624 hCoV-19/USA/WI-GMF-25868/2020|EPI_ISL_660937|2020-   ····················  ··C·················  ····················  ····················  ····················  ····················
S  6625 hCoV-19/USA/WI-GMF-25883/2020|EPI_ISL_660938|2020-   ····················  ····················  ····················  ····················  ····················  ····················
S  6626 hCoV-19/USA/IA-GMF-25923/2020|EPI_ISL_660939|2020-   ····················  ····················  ····················  ····················  ····················  ····················
S  6627 hCoV-19/USA/WI-GMF-25942/2020|EPI_ISL_660940|2020-   ····················  ····················  ····················  ····················  ····················  ····················
S  6628 hCoV-19/USA/WI-GMF-25948/2020|EPI_ISL_660941|2020-   ····················  ····················  ····················  ····················  ····················  ····················
S  6629 hCoV-19/USA/IA-GMF-25958/2020|EPI_ISL_660942|2020-   ····················  ····················  ····················  ····················  ····················  ····················
S  6630 hCoV-19/USA/MN-GMF-26024/2020|EPI_ISL_660943|2020-   ····················  ····················  ····················  ····················  ····················  ····················
S  6631 hCoV-19/USA/WI-GMF-26050/2020|EPI_ISL_660944|2020-   ····················  ····················  ····················  ····················  ····················  ···············A····
S  6632 hCoV-19/USA/WI-GMF-26052/2020|EPI_ISL_660945|2020-   ····················  ····················  ····················  ····················  ····················  ···············A····
S  6633 hCoV-19/USA/WI-GMF-26127/2020|EPI_ISL_660946|2020-   ····················  ····················  ····················  ····················  ····················  ····················
S  6634 hCoV-19/USA/WI-GMF-26151/2020|EPI_ISL_660947|2020-   ····················  ····················  ····················  ····················  ····················  ····················
S  6635 hCoV-19/USA/WI-GMF-26180/2020|EPI_ISL_660948|2020-   ····················  ····················  ····················  ····················  ····················  ····················
S  6636 hCoV-19/USA/WI-GMF-26332/2020|EPI_ISL_660949|2020-   ····················  ····················  ····················  ····················  ····················  ····················
S  6637 hCoV-19/USA/WI-GMF-26367/2020|EPI_ISL_660950|2020-   ····················  ····················  ····················  ····················  ····················  ····················
S  6638 hCoV-19/USA/WI-GMF-26474/2020|EPI_ISL_660951|2020-   ····················  ····················  ····················  ····················  ····················  ····················
S  6639 hCoV-19/USA/WI-GMF-26493/2020|EPI_ISL_660952|2020-   ····················  ····················  ····················  ····················  ····················  ····················
S  6640 hCoV-19/USA/WI-GMF-26494/2020|EPI_ISL_660953|2020-   ····················  ····················  ····················  ····················  ····················  ····················
S  6641 hCoV-19/USA/WI-GMF-26498/2020|EPI_ISL_660954|2020-   ····················  ····················  ····················  ····················  ····················  ····················
S  6642 hCoV-19/USA/WI-GMF-26517/2020|EPI_ISL_660955|2020-   ····················  ····················  ····················  ····················  ····················  ····················
S  6643 hCoV-19/USA/WI-GMF-26648/2020|EPI_ISL_660956|2020-   ····················  ····················  ····················  ····················  ····················  ····················
S  6644 hCoV-19/USA/WI-GMF-26654/2020|EPI_ISL_660957|2020-   ····················  ····················  ····················  ····················  ····················  ····················
S  6645 hCoV-19/USA/WI-GMF-26667/2020|EPI_ISL_660958|2020-   ····················  ····················  ····················  ····················  ····················  ····················
S  6646 hCoV-19/USA/WI-GMF-26669/2020|EPI_ISL_660959|2020-   ·······T············  ·A··················  ····················  ····················  ····················  ····················
S  6647 hCoV-19/USA/WI-GMF-26670/2020|EPI_ISL_660960|2020-   ····················  ····················  ····················  ····················  ····················  ···············A····
S  6648 hCoV-19/USA/WI-GMF-26682/2020|EPI_ISL_660961|2020-   ····················  ····················  ····················  ····················  ····················  ····················
S  6649 hCoV-19/USA/WI-GMF-26767/2020|EPI_ISL_660962|2020-   ····················  ·A··················  ····················  ····················  ····················  ····················
S  6650 hCoV-19/USA/WI-GMF-26859/2020|EPI_ISL_660963|2020-   ····················  ··C·················  ····················  ····················  ····················  ····················
S  6651 hCoV-19/USA/IA-GMF-26895/2020|EPI_ISL_660964|2020-   ····················  ·A··················  ····················  ····················  ····················  ····················
S  6652 hCoV-19/USA/WI-GMF-26896/2020|EPI_ISL_660965|2020-   ····················  ····················  ····················  ····················  ····················  ····················
S  6653 hCoV-19/USA/WI-GMF-27011/2020|EPI_ISL_660966|2020-   ····················  ····················  ····················  ····················  ····················  ····················
S  6654 hCoV-19/USA/MN-GMF-27046/2020|EPI_ISL_660967|2020-   ····················  ·A··················  ····················  ····················  ····················  ····················
S  6655 hCoV-19/USA/WI-GMF-27069/2020|EPI_ISL_660968|2020-   ····················  ····················  ····················  ····················  ····················  ····················
S  6656 hCoV-19/USA/WI-GMF-27154/2020|EPI_ISL_660969|2020-   ····················  ····················  ····················  ····················  ····················  ····················
S  6657 hCoV-19/Scotland/QEUH-B135A6/2020|EPI_ISL_649608|2   ····················  ····················  ····················  ····················  ····················  ····················
S  6658 hCoV-19/Scotland/QEUH-B1297E/2020|EPI_ISL_649610|2   ····················  ····················  ····················  ····················  ····················  ····················
S  6659 hCoV-19/Scotland/QEUH-B10F32/2020|EPI_ISL_649611|2   ····················  ····················  ····················  ····················  ····················  ····················
S  6660 hCoV-19/Scotland/QEUH-B13333/2020|EPI_ISL_649612|2   ····················  ····················  ····················  ····················  ····················  ····················
S  6661 hCoV-19/Scotland/QEUH-B132DC/2020|EPI_ISL_649613|2   ····················  ····················  ····················  ····················  ····················  ····················
S  6662 hCoV-19/Scotland/QEUH-B1337F/2020|EPI_ISL_649614|2   ····················  ····················  ····················  ····················  ····················  ····················
S  6663 hCoV-19/Scotland/QEUH-B1299C/2020|EPI_ISL_649615|2   ····················  ····················  ····················  ····················  ····················  ····················
S  6664 hCoV-19/Scotland/QEUH-B1339D/2020|EPI_ISL_649616|2   ····················  ····················  ····················  ····················  ····················  ····················
S  6665 hCoV-19/Scotland/QEUH-B135D3/2020|EPI_ISL_649617|2   ····················  ····················  ····················  ····················  ····················  ····················
S  6666 hCoV-19/Scotland/QEUH-B10BA7/2020|EPI_ISL_649618|2   ····················  ····················  ····················  ····················  ····················  ····················
S  6667 hCoV-19/Scotland/QEUH-B1261D/2020|EPI_ISL_649619|2   ····················  ····················  ····················  ····················  ····················  ····················
S  6668 hCoV-19/Scotland/QEUH-B108EC/2020|EPI_ISL_649620|2   ····················  ····················  ····················  ····················  ····················  ····················
S  6669 hCoV-19/Scotland/QEUH-B10A8C/2020|EPI_ISL_649621|2   ····················  ····················  ····················  ····················  ····················  ····················
```

| | | | | | | | |
|---|---|---|---|---|---|---|---|
| S | 6670 hCoV-19/Scotland/QEUH-B12589/2020\|EPI_ISL_649622\|2 | ···················· | ···················· | ····················· | ···················· | ···················· | ···················· |
| S | 6671 hCoV-19/Scotland/QEUH-B125B6/2020\|EPI_ISL_649624\|2 | ···················· | ···················· | ····················· | ···················· | ···················· | ···················· |
| S | 6672 hCoV-19/England/CAMC-B0802B/2020\|EPI_ISL_649625\|20 | ···················· | ···················· | ····················· | ···················· | ···················· | ···················· |
| S | 6673 hCoV-19/England/CAMC-B0C670/2020\|EPI_ISL_649626\|20 | ···················· | ···················· | ····················· | ···················· | ···················· | ···················· |
| S | 6674 hCoV-19/England/CAMC-B0C661/2020\|EPI_ISL_649627\|20 | ···················· | ···················· | ····················· | ···················· | ···················· | ···················· |
| S | 6675 hCoV-19/England/CAMC-B0C485/2020\|EPI_ISL_649628\|20 | ···················· | ···················· | ····················· | ···················· | ···················· | ···················· |
| S | 6676 hCoV-19/England/CAMC-B07F6B/2020\|EPI_ISL_649630\|20 | ···················· | ···················· | ····················· | ···················· | ······T············· | ···················· |
| S | 6677 hCoV-19/England/CAMC-B0829E/2020\|EPI_ISL_649631\|20 | ···················· | ···················· | ····················· | ···················· | ···················· | ···················· |
| S | 6678 hCoV-19/England/CAMC-B076B1/2020\|EPI_ISL_649632\|20 | ···················· | ···················· | ····················· | ···················· | ····T··············· | ···················· |
| S | 6679 hCoV-19/England/CAMC-B07499/2020\|EPI_ISL_649633\|20 | ···················· | ···················· | ····················· | ···················· | ···················· | ···················· |
| S | 6680 hCoV-19/Scotland/QEUH-AE2F2B/2020\|EPI_ISL_649634\|2 | ···················· | ···················· | ····················· | ···················· | ···················· | ···················· |
| S | 6681 hCoV-19/Scotland/QEUH-AE20EC/2020\|EPI_ISL_649635\|2 | ···················· | ···················· | ····················· | ···················· | ···················· | ···················· |
| S | 6682 hCoV-19/Scotland/QEUH-AE2E88/2020\|EPI_ISL_649636\|2 | ···················· | ···················· | ····················· | ···················· | ···················· | ···················· |
| S | 6683 hCoV-19/Scotland/QEUH-AE1DB9/2020\|EPI_ISL_649637\|2 | ···················· | ···················· | ····················· | ···················· | ···················· | ···················· |
| S | 6684 hCoV-19/Scotland/QEUH-AE19D3/2020\|EPI_ISL_649638\|2 | ···················· | ···················· | ····················· | ···················· | ···················· | ···················· |
| S | 6685 hCoV-19/Scotland/QEUH-AE2F85/2020\|EPI_ISL_649639\|2 | ···················· | ···················· | ····················· | ···················· | ···················· | ···················· |
| S | 6686 hCoV-19/Scotland/QEUH-AE1D7D/2020\|EPI_ISL_649640\|2 | ···················· | ···················· | ····················· | ···················· | ···················· | ···················· |
| S | 6687 hCoV-19/Scotland/QEUH-AE1F3B/2020\|EPI_ISL_649641\|2 | ···················· | ···················· | ····················· | ···················· | ···················· | ···················· |
| S | 6688 hCoV-19/Scotland/QEUH-AE1F4A/2020\|EPI_ISL_649642\|2 | ···················· | ···················· | ····················· | ···················· | ···················· | ···················· |
| S | 6689 hCoV-19/Scotland/QEUH-AE2EA6/2020\|EPI_ISL_649643\|2 | ···················· | ···················· | ····················· | ···················· | ···················· | ···················· |
| S | 6690 hCoV-19/Scotland/QEUH-AE2662/2020\|EPI_ISL_649644\|2 | ···················· | ···················· | ····················· | ···················· | ···················· | ···················· |
| S | 6691 hCoV-19/Scotland/QEUH-AE1F0E/2020\|EPI_ISL_649645\|2 | ···················· | ···················· | ····················· | ···················· | ···················· | ···················· |
| S | 6692 hCoV-19/Scotland/QEUH-AE28F3/2020\|EPI_ISL_649646\|2 | ···················· | ···················· | ····················· | ···················· | ···················· | ···················· |
| S | 6693 hCoV-19/Scotland/QEUH-AE2F49/2020\|EPI_ISL_649647\|2 | ···················· | ···················· | ····················· | ···················· | ···················· | ···················· |
| S | 6694 hCoV-19/Scotland/QEUH-AE1EE3/2020\|EPI_ISL_649648\|2 | ···················· | ···················· | ····················· | ···················· | ···················· | ···················· |
| S | 6695 hCoV-19/Scotland/QEUH-AE4B07/2020\|EPI_ISL_649661\|2 | ···················· | ···················· | ····················· | ···················· | ···················· | ···················· |
| S | 6696 hCoV-19/Scotland/QEUH-AE5456/2020\|EPI_ISL_649662\|2 | ···················· | ···················· | ····················· | ···················· | ···················· | ···················· |
| S | 6697 hCoV-19/Scotland/QEUH-AE4C8C/2020\|EPI_ISL_649663\|2 | ···················· | ···················· | ····················· | ···················· | ···················· | ···················· |
| S | 6698 hCoV-19/Scotland/QEUH-AE49D0/2020\|EPI_ISL_649666\|2 | ···················· | ···················· | ····················· | ···················· | ···················· | ···················· |
| S | 6699 hCoV-19/Scotland/QEUH-AE4A0A/2020\|EPI_ISL_649667\|2 | ···················· | ···················· | ····················· | ···················· | ···················· | ···················· |
| S | 6700 hCoV-19/Scotland/QEUH-AE5113/2020\|EPI_ISL_649668\|2 | ···················· | ···················· | ····················· | ···················· | ···················· | ···················· |
| S | 6701 hCoV-19/Scotland/QEUH-AE4BCB/2020\|EPI_ISL_649669\|2 | ···················· | ···················· | ····················· | ···················· | ···················· | ···················· |
| S | 6702 hCoV-19/Scotland/QEUH-AD53A7/2020\|EPI_ISL_649690\|2 | ···················· | ···················· | ····················· | ···················· | ···················· | ···················· |
| S | 6703 hCoV-19/Scotland/QEUH-AD150F/2020\|EPI_ISL_649691\|2 | ···················· | ···················· | ····················· | ···················· | ···················· | ···················· |
| S | 6704 hCoV-19/Scotland/QEUH-AD53D4/2020\|EPI_ISL_649692\|2 | ···················· | ···················· | ····················· | ···················· | ···················· | ···················· |
| S | 6705 hCoV-19/Scotland/QEUH-AD436C/2020\|EPI_ISL_649693\|2 | ···················· | ···················· | ····················· | ···················· | ···················· | ···················· |
| S | 6706 hCoV-19/Scotland/QEUH-AD144E/2020\|EPI_ISL_649694\|2 | ···················· | ···················· | ····················· | ···················· | ···················· | ···················· |
| S | 6707 hCoV-19/Scotland/QEUH-AD5B36/2020\|EPI_ISL_649695\|2 | ···················· | ···················· | ····················· | ···················· | ···················· | ···················· |
| S | 6708 hCoV-19/Scotland/QEUH-AD541D/2020\|EPI_ISL_649696\|2 | ···················· | ···················· | ····················· | ···················· | ···················· | ···················· |
| S | 6709 hCoV-19/Scotland/QEUH-AD3FF0/2020\|EPI_ISL_649697\|2 | ···················· | ···················· | ····················· | ···················· | ···················· | ···················· |
| S | 6710 hCoV-19/Scotland/QEUH-AD18F7/2020\|EPI_ISL_649698\|2 | ···················· | ···················· | ····················· | ···················· | ···················· | ···················· |
| S | 6711 hCoV-19/Scotland/QEUH-AD146C/2020\|EPI_ISL_649699\|2 | ···················· | ···················· | ····················· | ···················· | ···················· | ···················· |
| S | 6712 hCoV-19/Scotland/CVR5701/2020\|EPI_ISL_651920\|2020- | ···················· | ···················· | ····················· | ···················· | ···················· | ···················· |
| S | 6713 hCoV-19/Scotland/CVR5702/2020\|EPI_ISL_651921\|2020- | ···················· | ···················· | ····················· | ···················· | ···················· | ···················· |
| S | 6714 hCoV-19/Scotland/CVR5704/2020\|EPI_ISL_651922\|2020- | ···················· | ···················· | ····················· | ···················· | ···················· | ···················· |
| S | 6715 hCoV-19/Scotland/CVR5705/2020\|EPI_ISL_651923\|2020- | ···················· | ···················· | ····················· | ···················· | ···················· | ···················· |
| S | 6716 hCoV-19/Scotland/CVR5706/2020\|EPI_ISL_651924\|2020- | ···················· | ···················· | ····················· | ···················· | ···················· | ···················· |
| S | 6717 hCoV-19/Scotland/CVR5707/2020\|EPI_ISL_651925\|2020- | ···················· | ···················· | ····················· | ···················· | ···················· | ···················· |
| S | 6718 hCoV-19/Scotland/CVR5708/2020\|EPI_ISL_651926\|2020- | ···················· | ···················· | ····················· | ···················· | ···················· | ···················· |
| S | 6719 hCoV-19/Scotland/CVR5709/2020\|EPI_ISL_651927\|2020- | ···················· | ···················· | ····················· | ···················· | ···················· | ···················· |
| S | 6720 hCoV-19/Scotland/CVR5711/2020\|EPI_ISL_651928\|2020- | ···················· | ···················· | ····················· | ···················· | ···················· | ···················· |
| S | 6721 hCoV-19/Scotland/CVR5713/2020\|EPI_ISL_651929\|2020- | ···················· | ···················· | ····················· | ···················· | ···················· | ···················· |
| S | 6722 hCoV-19/Scotland/CVR5716/2020\|EPI_ISL_651930\|2020- | ···················· | ···················· | ····················· | ···················· | ···················· | ···················· |
| S | 6723 hCoV-19/Scotland/CVR5717/2020\|EPI_ISL_651931\|2020- | ···················· | ·A·················· | ····················· | ···················· | ···················· | ···················· |
| S | 6724 hCoV-19/Scotland/CVR5719/2020\|EPI_ISL_651932\|2020- | ···················· | ···················· | ····················· | ···················· | ···················· | ···················· |
| S | 6725 hCoV-19/Scotland/CVR5720/2020\|EPI_ISL_651933\|2020- | ···················· | ···················· | ····················· | ···················· | ···················· | ···················· |
| S | 6726 hCoV-19/Scotland/CVR5721/2020\|EPI_ISL_651934\|2020- | ···················· | ···················· | ····················· | ···················· | ···················· | ···················· |
| S | 6727 hCoV-19/Scotland/CVR5724/2020\|EPI_ISL_651935\|2020- | ···················· | ···················· | ····················· | ···················· | ···················· | ···················· |
| S | 6728 hCoV-19/Scotland/CVR5725/2020\|EPI_ISL_651936\|2020- | ···················· | ···················· | ····················· | ···················· | ···················· | ···················· |
| S | 6729 hCoV-19/Scotland/CVR5726/2020\|EPI_ISL_651937\|2020- | ···················· | ···················· | ····················· | ···················· | ···················· | ···················· |
| S | 6730 hCoV-19/Scotland/CVR5728/2020\|EPI_ISL_651938\|2020- | ···················· | ···················· | ····················· | ···················· | ···················· | ···················· |
| S | 6731 hCoV-19/Scotland/CVR5729/2020\|EPI_ISL_651939\|2020- | ···················· | ···················· | ····················· | ···················· | ···················· | ···················· |
| S | 6732 hCoV-19/Scotland/CVR5730/2020\|EPI_ISL_651940\|2020- | ···················· | ···················· | ····················· | ···················· | ···················· | ···················· |
| S | 6733 hCoV-19/Scotland/CVR5731/2020\|EPI_ISL_651941\|2020- | ···················· | ···················· | ····················· | ···················· | ···················· | ···················· |
| S | 6734 hCoV-19/Scotland/CVR5732/2020\|EPI_ISL_651942\|2020- | ···················· | ···················· | ····················· | ···················· | ···················· | ···················· |
| S | 6735 hCoV-19/Scotland/CVR5733/2020\|EPI_ISL_651943\|2020- | ···················· | ···················· | ····················· | ···················· | ···················· | ···················· |
| S | 6736 hCoV-19/Scotland/CVR5734/2020\|EPI_ISL_651944\|2020- | ···················· | ···················· | ····················· | ···················· | ···················· | ···················· |
| S | 6737 hCoV-19/Scotland/CVR5736/2020\|EPI_ISL_651945\|2020- | ···················· | ···················· | ····················· | ···················· | ···················· | ···················· |
| S | 6738 hCoV-19/Scotland/CVR5738/2020\|EPI_ISL_651946\|2020- | ···················· | ···················· | ····················· | ···················· | ···················· | ···················· |
| S | 6739 hCoV-19/Scotland/CVR5740/2020\|EPI_ISL_651947\|2020- | ···················· | ···················· | ····················· | ···················· | ···················· | ···················· |
| S | 6740 hCoV-19/Scotland/CVR5741/2020\|EPI_ISL_651948\|2020- | ···················· | ···················· | ····················· | ···················· | ···················· | ···················· |
| S | 6741 hCoV-19/Scotland/CVR5743/2020\|EPI_ISL_651949\|2020- | ···················· | ···················· | ····················· | ···················· | ···················· | ···················· |
| S | 6742 hCoV-19/Scotland/CVR5744/2020\|EPI_ISL_651950\|2020- | ···················· | ···················· | ····················· | ···················· | ···················· | ···················· |
| S | 6743 hCoV-19/Scotland/CVR5745/2020\|EPI_ISL_651951\|2020- | ···················· | ···················· | ····················· | ···················· | ···················· | ···················· |
| S | 6744 hCoV-19/Scotland/CVR5746/2020\|EPI_ISL_651952\|2020- | ···················· | ···················· | ····················· | ···················· | ···················· | ···················· |
| S | 6745 hCoV-19/Scotland/CVR5747/2020\|EPI_ISL_651953\|2020- | ···················· | ···················· | ····················· | ···················· | ···················· | ···················· |
| S | 6746 hCoV-19/Scotland/CVR5749/2020\|EPI_ISL_651954\|2020- | ···················· | ···················· | ····················· | ···················· | ···················· | ···················· |
| S | 6747 hCoV-19/Scotland/CVR5750/2020\|EPI_ISL_651955\|2020- | ···················· | ···················· | ····················· | ···················· | ···················· | ···················· |
| S | 6748 hCoV-19/Scotland/CVR5751/2020\|EPI_ISL_651956\|2020- | ···················· | ···················· | ····················· | ···················· | ···················· | ···················· |
| S | 6749 hCoV-19/Scotland/EDB8058/2020\|EPI_ISL_651957\|2020- | ···················· | ···················· | ····················· | ···················· | ···················· | ···················· |

| | | | | | | | |
|---|---|---|---|---|---|---|---|
| S | 6750 hCoV-19/Scotland/EDB8481/2020\|EPI_ISL_651958\|2020- | .................... | .................... | .................... | .................... | .................... | .................... |
| S | 6751 hCoV-19/Scotland/EDB8483/2020\|EPI_ISL_651959\|2020- | .................... | .................... | .................... | .................... | .................... | .................... |
| S | 6752 hCoV-19/Scotland/EDB8564/2020\|EPI_ISL_651960\|2020- | .................... | .................... | .................... | .................... | .................... | .................... |
| S | 6753 hCoV-19/Scotland/EDB8592/2020\|EPI_ISL_651972\|2020- | .................... | .................... | .................... | .................... | .................... | .................... |
| S | 6754 hCoV-19/Scotland/EDB8593/2020\|EPI_ISL_651973\|2020- | .................... | .................... | .................... | .................... | .................... | .................... |
| S | 6755 hCoV-19/Scotland/EDB8601/2020\|EPI_ISL_651976\|2020- | .................... | .................... | .................... | .................... | .................... | .................... |
| S | 6756 hCoV-19/Scotland/EDB8602/2020\|EPI_ISL_651977\|2020- | .................... | .................... | .................... | .................... | .................... | .................... |
| S | 6757 hCoV-19/Scotland/EDB8603/2020\|EPI_ISL_651978\|2020- | .................... | .................... | .................... | .................... | .................... | .................... |
| S | 6758 hCoV-19/Scotland/EDB8609/2020\|EPI_ISL_651979\|2020- | .................... | .................... | .................... | .................... | .................... | .................... |
| S | 6762 hCoV-19/Spain/MD-IBV-99008563/2020\|EPI_ISL_654103\| | .................... | .................... | .................... | .................... | .................... | .................... |
| S | 6763 hCoV-19/Spain/MD-IBV-99008564/2020\|EPI_ISL_654104\| | .................... | .................... | .................... | .................... | .................... | .................... |
| S | 6765 hCoV-19/England/ALDP-B62F13/2020\|EPI_ISL_655428\|20 | .................... | .................... | .................... | .................... | .................... | .................... |
| S | 6766 hCoV-19/England/ALDP-B63D54/2020\|EPI_ISL_655430\|20 | .................... | .................... | .................... | .................... | .................... | .................... |
| S | 6767 hCoV-19/England/ALDP-B5B469/2020\|EPI_ISL_655432\|20 | .................... | .................... | .................... | .................... | .................... | .................... |
| S | 6768 hCoV-19/England/ALDP-B5B5A2/2020\|EPI_ISL_655433\|20 | .................... | .................... | .................... | .................... | .................... | .................... |
| S | 6769 hCoV-19/England/ALDP-B63C0C/2020\|EPI_ISL_655434\|20 | .................... | .................... | .................... | .................... | .................... | .................... |
| S | 6770 hCoV-19/England/ALDP-B5B1EA/2020\|EPI_ISL_655435\|20 | .................... | .................... | .................... | .................... | .................... | .................... |
| S | 6771 hCoV-19/England/CAMC-B52338/2020\|EPI_ISL_656760\|20 | .................... | .................... | .................... | .................... | .................... | .................... |
| S | 6772 hCoV-19/England/CAMC-B54002/2020\|EPI_ISL_656761\|20 | .................... | .A.................. | .................... | .................... | .................... | ..A................. |
| S | 6773 hCoV-19/England/CAMC-B543BE/2020\|EPI_ISL_656762\|20 | .................... | .................... | .................... | .................... | .................... | .................... |
| S | 6774 hCoV-19/England/CAMC-B53F7F/2020\|EPI_ISL_656764\|20 | .................... | .................... | .................... | .................... | .................... | .................... |
| S | 6775 hCoV-19/England/CAMC-B52550/2020\|EPI_ISL_656766\|20 | .................... | .................... | .................... | .................... | .................... | .................... |
| S | 6776 hCoV-19/England/CAMC-B542C0/2020\|EPI_ISL_656767\|20 | .................... | .................... | .................... | .................... | .................... | .................... |
| S | 6777 hCoV-19/England/CAMC-B522E0/2020\|EPI_ISL_656800\|20 | .................... | .................... | .................... | .................... | .................... | .................... |
| S | 6778 hCoV-19/England/CAMC-B544AC/2020\|EPI_ISL_656802\|20 | .................... | .................... | .................... | .................... | .................... | .................... |
| S | 6779 hCoV-19/England/CAMC-B53759/2020\|EPI_ISL_656803\|20 | .................... | .................... | .................... | .................... | .................... | .................... |
| S | 6780 hCoV-19/England/CAMC-B54327/2020\|EPI_ISL_656804\|20 | .................... | .................... | .................... | .................... | .................... | .................... |
| S | 6781 hCoV-19/England/CAMC-B5222C/2020\|EPI_ISL_656805\|20 | .................... | .................... | .................... | .................... | .................... | .................... |
| S | 6782 hCoV-19/England/CAMC-B52435/2020\|EPI_ISL_656806\|20 | .................... | .................... | .................... | .................... | .................... | .................... |
| S | 6783 hCoV-19/England/MILK-B56C84/2020\|EPI_ISL_656807\|20 | .................... | .................... | .................... | .................... | .................... | .................... |
| S | 6784 hCoV-19/England/MILK-B56A02/2020\|EPI_ISL_656808\|20 | .................... | .................... | .................... | .................... | .................... | .................... |
| S | 6785 hCoV-19/England/CAMC-B5221D/2020\|EPI_ISL_656809\|20 | .................... | .A.................. | .................... | .................... | .................... | ..A................. |
| S | 6786 hCoV-19/England/MILK-B56BD2/2020\|EPI_ISL_656810\|20 | .................... | .................... | .................... | .................... | .................... | .................... |
| S | 6787 hCoV-19/England/MILK-B56792/2020\|EPI_ISL_656811\|20 | .................... | .................... | .................... | .................... | .................... | .................... |
| S | 6788 hCoV-19/England/MILK-B3866C/2020\|EPI_ISL_659010\|20 | .................... | .................... | .................... | .................... | .................... | .................... |
| S | 6789 hCoV-19/England/CAMC-B37F9F/2020\|EPI_ISL_659011\|20 | .................... | .................... | .................... | .................... | .................... | .................... |
| S | 6790 hCoV-19/England/MILK-B387A5/2020\|EPI_ISL_659012\|20 | .................... | .................... | .................... | .................... | .................... | .................... |
| S | 6791 hCoV-19/England/MILK-B395C8/2020\|EPI_ISL_659014\|20 | .................... | .................... | .................... | .................... | .................... | .................... |
| S | 6792 hCoV-19/England/CAMC-B37F44/2020\|EPI_ISL_659017\|20 | .................... | .................... | .................... | .................... | .................... | .................... |
| S | 6793 hCoV-19/England/CAMC-B37D86/2020\|EPI_ISL_659018\|20 | .................... | .................... | .................... | .................... | .................... | .................... |
| S | 6794 hCoV-19/England/CAMC-B38022/2020\|EPI_ISL_659019\|20 | .................... | .................... | .................... | .................... | .................... | .................... |
| S | 6795 hCoV-19/England/MILK-B38D2B/2020\|EPI_ISL_659020\|20 | .................... | .................... | .................... | .................... | .................... | .................... |
| S | 6796 hCoV-19/England/MILK-B3A521/2020\|EPI_ISL_659023\|20 | .................... | .................... | .................... | .................... | .................... | .................... |
| S | 6797 hCoV-19/England/MILK-B38AE8/2020\|EPI_ISL_659024\|20 | .................... | .................... | .................... | .................... | .................... | .................... |
| S | 6798 hCoV-19/England/MILK-B3957D/2020\|EPI_ISL_659026\|20 | .................... | .................... | .................... | .................... | .................... | .................... |
| S | 6799 hCoV-19/England/MILK-B384F9/2020\|EPI_ISL_659027\|20 | .................... | .................... | .................... | .................... | .................... | .................... |
| S | 6800 hCoV-19/England/CAMC-B37DD1/2020\|EPI_ISL_659028\|20 | .................... | .................... | .................... | .................... | .................... | .................... |
| S | 6801 hCoV-19/England/MILK-B38796/2020\|EPI_ISL_659029\|20 | .................... | .A.................. | .................... | .................... | .................... | ..A................. |
| S | 6802 hCoV-19/England/MILK-B3914C/2020\|EPI_ISL_659030\|20 | .................... | .................... | .................... | .................... | .................... | ...............R.... |
| S | 6803 hCoV-19/England/MILK-B3859C/2020\|EPI_ISL_659031\|20 | .................... | .A.................. | .................... | .................... | .................... | ..A................. |
| S | 6804 hCoV-19/England/MILK-B3A5A9/2020\|EPI_ISL_659032\|20 | .................... | .................... | .................... | .................... | .................... | .................... |
| S | 6805 hCoV-19/England/MILK-B395F5/2020\|EPI_ISL_659033\|20 | .................... | .................... | .................... | .................... | .................... | .................... |
| S | 6806 hCoV-19/England/MILK-B3A530/2020\|EPI_ISL_659034\|20 | .................... | .................... | .................... | .................... | .................... | .................... |
| S | 6807 hCoV-19/England/MILK-B3857E/2020\|EPI_ISL_659036\|20 | .................... | .................... | .................... | .................... | .................... | .................... |
| S | 6808 hCoV-19/England/MILK-B3A62E/2020\|EPI_ISL_659037\|20 | .................... | .................... | .................... | .................... | .................... | .................... |
| S | 6809 hCoV-19/England/MILK-B38C00/2020\|EPI_ISL_659038\|20 | .................... | .................... | .................... | .................... | .................... | .................... |
| S | 6810 hCoV-19/England/MILK-B38839/2020\|EPI_ISL_659039\|20 | .................... | .A.................. | .................... | .................... | .................... | ..A................. |
| S | 6811 hCoV-19/England/MILK-B38541/2020\|EPI_ISL_659042\|20 | .................... | .................... | .................... | .................... | .................... | .................... |
| S | 6812 hCoV-19/England/MILK-B384EA/2020\|EPI_ISL_659043\|20 | .................... | .A.................. | .................... | .................... | .................... | ..A................. |
| S | 6813 hCoV-19/England/MILK-B389AF/2020\|EPI_ISL_659044\|20 | .................... | .................... | .................... | .................... | .................... | .................... |
| S | 6814 hCoV-19/England/CAMC-B37D3B/2020\|EPI_ISL_659045\|20 | .................... | .A.................. | .................... | .................... | .................... | ..A................. |
| S | 6815 hCoV-19/England/MILK-B38471/2020\|EPI_ISL_659047\|20 | .................... | .................... | .................... | .................... | .................... | .................... |
| S | 6816 hCoV-19/England/MILK-B38A8E/2020\|EPI_ISL_659049\|20 | .................... | .................... | .................... | .................... | .................... | .................... |
| S | 6817 hCoV-19/England/CAMC-B37FEA/2020\|EPI_ISL_659050\|20 | .................... | .................... | .................... | .................... | .................... | .................... |
| S | 6818 hCoV-19/England/MILK-B38D67/2020\|EPI_ISL_659051\|20 | .................... | .................... | .................... | .................... | .................... | .................... |
| S | 6819 hCoV-19/England/CAMC-B38040/2020\|EPI_ISL_659053\|20 | .................... | .................... | .................... | .................... | ......T............. | .................... |
| S | 6820 hCoV-19/England/CAMC-B37CA7/2020\|EPI_ISL_659054\|20 | .................... | .................... | .................... | .................... | .................... | .................... |
| S | 6821 hCoV-19/England/MILK-B38E73/2020\|EPI_ISL_659055\|20 | .................... | .................... | .................... | .................... | .................... | .................... |
| S | 6822 hCoV-19/England/MILK-B3A54F/2020\|EPI_ISL_659056\|20 | .................... | .................... | .................... | .................... | .................... | .................... |
| S | 6823 hCoV-19/England/MILK-B38D58/2020\|EPI_ISL_659058\|20 | .................... | .................... | .................... | .................... | .................... | .................... |
| S | 6824 hCoV-19/England/MILK-B38AF7/2020\|EPI_ISL_659060\|20 | .................... | .................... | .................... | .................... | .................... | .................... |
| S | 6825 hCoV-19/England/MILK-B38CD3/2020\|EPI_ISL_659061\|20 | .................... | .................... | .................... | .................... | .................... | .................... |
| S | 6826 hCoV-19/England/MILK-B38523/2020\|EPI_ISL_659062\|20 | .................... | .................... | .................... | .................... | .................... | .................... |
| S | 6827 hCoV-19/England/MILK-B38CC4/2020\|EPI_ISL_659063\|20 | .................... | .................... | .................... | .................... | .................... | .................... |
| S | 6828 hCoV-19/England/MILK-B393DD/2020\|EPI_ISL_659064\|20 | .................... | .................... | .................... | .................... | .................... | .................... |
| S | 6829 hCoV-19/England/MILK-B39373/2020\|EPI_ISL_659065\|20 | .................... | .................... | .................... | .................... | .................... | .................... |
| S | 6830 hCoV-19/England/MILK-B38875/2020\|EPI_ISL_659066\|20 | .................... | .................... | .................... | .................... | .................... | .................... |
| S | 6831 hCoV-19/England/MILK-B388DF/2020\|EPI_ISL_659068\|20 | .................... | .A.................. | .................... | .................... | .................... | ..A................. |
| S | 6832 hCoV-19/England/MILK-B38787/2020\|EPI_ISL_659069\|20 | .................... | .................... | .................... | .................... | .................... | .................... |
| S | 6833 hCoV-19/England/CAMC-B3815C/2020\|EPI_ISL_659071\|20 | .................... | .................... | .................... | .................... | .................... | .................... |

| | | | | | | | | |
|---|---|---|---|---|---|---|---|---|
| S | 6834 | hCoV-19/England/CAMC-B37DC2/2020\|EPI_ISL_659072\|20 | ···················· | ···················· | ····················· | ···················· | ···················· | ···················· |
| S | 6835 | hCoV-19/England/CAMC-B382E0/2020\|EPI_ISL_659073\|20 | ···················· | ···················· | ····················· | ···················· | ···················· | ···················· |
| S | 6836 | hCoV-19/England/MILK-B38505/2020\|EPI_ISL_659074\|20 | ···················· | ···················· | ····················· | ···················· | ···················· | ···················· |
| S | 6837 | hCoV-19/England/MILK-B39197/2020\|EPI_ISL_659075\|20 | ···················· | ···················· | ····················· | ···················· | ···················· | ···················· |
| S | 6838 | hCoV-19/England/CAMC-B38268/2020\|EPI_ISL_659076\|20 | ···················· | ···················· | ····················· | ···················· | ···················· | ···················· |
| S | 6839 | hCoV-19/England/CAMC-B37D2C/2020\|EPI_ISL_659077\|20 | ···················· | ···················· | ····················· | ···················· | ···················· | ···················· |
| S | 6840 | hCoV-19/England/MILK-B38D49/2020\|EPI_ISL_659078\|20 | ···················· | ···················· | ····················· | ···················· | ···················· | ···················· |
| S | 6841 | hCoV-19/England/CAMC-B37EED/2020\|EPI_ISL_659080\|20 | ···················· | ···················· | ····················· | ···················· | ···················· | ···················· |
| S | 6842 | hCoV-19/England/MILK-B38866/2020\|EPI_ISL_659081\|20 | ···················· | ···················· | ····················· | ···················· | ···················· | ···················· |
| S | 6843 | hCoV-19/England/CAMC-B37E56/2020\|EPI_ISL_659083\|20 | ···················· | ···················· | ····················· | ···················· | ···················· | ···················· |
| S | 6844 | hCoV-19/England/CAMC-B37FAE/2020\|EPI_ISL_659084\|20 | ···················· | ···················· | ····················· | ···················· | ···················· | ···················· |
| S | 6845 | hCoV-19/England/MILK-B3A1E1/2020\|EPI_ISL_659085\|20 | ···················· | ···················· | ····················· | ···················· | ···················· | ···················· |
| S | 6846 | hCoV-19/England/MILK-B38E28/2020\|EPI_ISL_659086\|20 | ···················· | ···················· | ····················· | ···················· | ···················· | ···················· |
| S | 6847 | hCoV-19/England/MILK-B389DC/2020\|EPI_ISL_659087\|20 | ···················· | ···················· | ····················· | ···················· | ···················· | ···················· |
| S | 6848 | hCoV-19/England/MILK-B38A51/2020\|EPI_ISL_659089\|20 | ···················· | ···················· | ····················· | ···················· | ···················· | ···················· |
| S | 6849 | hCoV-19/England/QEUH-B2E827/2020\|EPI_ISL_660000\|20 | ···················· | ···················· | ····················· | ···················· | ···················· | ···················· |
| S | 6850 | hCoV-19/England/QEUH-B2F64A/2020\|EPI_ISL_660001\|20 | ···················· | ···················· | ····················· | ···················· | ···················· | ···················· |
| S | 6851 | hCoV-19/England/QEUH-B2D8A0/2020\|EPI_ISL_660005\|20 | ···················· | ···················· | ····················· | ···················· | ···················· | ···················· |
| S | 6852 | hCoV-19/England/QEUH-B2CEAF/2020\|EPI_ISL_660006\|20 | ···················· | ···················· | ····················· | ···················· | ···················· | ···················· |
| S | 6853 | hCoV-19/England/QEUH-B30B29/2020\|EPI_ISL_660007\|20 | ···················· | ···················· | ····················· | ···················· | ······T············· | ···················· |
| S | 6854 | hCoV-19/England/QEUH-B2D4AC/2020\|EPI_ISL_660008\|20 | ···················· | ···················· | ····················· | ···················· | ···················· | ···················· |
| S | 6855 | hCoV-19/England/QEUH-B2E97F/2020\|EPI_ISL_660010\|20 | ···················· | ···················· | ····················· | ···················· | ···················· | ···················· |
| S | 6856 | hCoV-19/England/QEUH-B2EB97/2020\|EPI_ISL_660011\|20 | ···················· | ···················· | ····················· | ···················· | ···················· | ···················· |
| S | 6857 | hCoV-19/England/QEUH-B2D4E8/2020\|EPI_ISL_660013\|20 | ···················· | ···················· | ····················· | ···················· | ···················· | ···················· |
| S | 6858 | hCoV-19/England/QEUH-B2DECC/2020\|EPI_ISL_660014\|20 | ···················· | ···················· | ····················· | ···················· | ···················· | ···················· |
| S | 6859 | hCoV-19/England/QEUH-B2D2EE/2020\|EPI_ISL_660015\|20 | ···················· | ···················· | ····················· | ···················· | ···················· | ···················· |
| S | 6860 | hCoV-19/England/QEUH-B2DEAE/2020\|EPI_ISL_660016\|20 | ···················· | ···················· | ····················· | ···················· | ···················· | ···················· |
| S | 6861 | hCoV-19/England/QEUH-B2D2DF/2020\|EPI_ISL_660019\|20 | ···················· | ···················· | ····················· | ···················· | ···················· | ···················· |
| S | 6862 | hCoV-19/England/QEUH-B2CD2A/2020\|EPI_ISL_660020\|20 | ···················· | ···················· | ····················· | ···················· | ···················· | ···················· |
| S | 6863 | hCoV-19/England/QEUH-B2EEE9/2020\|EPI_ISL_660021\|20 | ···················· | ···················· | ····················· | ···················· | ···················· | ···················· |
| S | 6864 | hCoV-19/England/QEUH-B3036D/2020\|EPI_ISL_660022\|20 | ···················· | ···················· | ····················· | ···················· | ···················· | ···················· |
| S | 6865 | hCoV-19/England/QEUH-B2F659/2020\|EPI_ISL_660023\|20 | ···················· | ···················· | ····················· | ···················· | ···················· | ···················· |
| S | 6866 | hCoV-19/England/QEUH-B2F6E0/2020\|EPI_ISL_660024\|20 | ···················· | ···················· | ····················· | ···················· | ···················· | ···················· |
| S | 6867 | hCoV-19/England/QEUH-B2DFBA/2020\|EPI_ISL_660025\|20 | ···················· | ···················· | ····················· | ···················· | ···················· | ···················· |
| S | 6868 | hCoV-19/England/QEUH-B2D293/2020\|EPI_ISL_660027\|20 | ···················· | ···················· | ····················· | ···················· | ···················· | ···················· |
| S | 6869 | hCoV-19/England/QEUH-B2CCF0/2020\|EPI_ISL_660028\|20 | ···················· | ···················· | ·················Y··· | ···················· | ···················· | ···················· |
| S | 6870 | hCoV-19/England/QEUH-B2EC2B/2020\|EPI_ISL_660029\|20 | ···················· | ···················· | ····················· | ···················· | ···················· | ···················· |
| S | 6871 | hCoV-19/England/QEUH-B2E8AF/2020\|EPI_ISL_660030\|20 | ···················· | ···················· | ····················· | ···················· | ···················· | ···················· |
| S | 6872 | hCoV-19/England/QEUH-B30BCF/2020\|EPI_ISL_660032\|20 | ···················· | ···················· | ····················· | ···················· | ···················· | ···················· |
| S | 6873 | hCoV-19/England/QEUH-B30BB0/2020\|EPI_ISL_660033\|20 | ···················· | ···················· | ····················· | ···················· | ···················· | ···················· |
| S | 6874 | hCoV-19/England/QEUH-B2D318/2020\|EPI_ISL_660034\|20 | ···················· | ···················· | ····················· | ···················· | ···················· | ···················· |
| S | 6875 | hCoV-19/England/QEUH-B2EF7D/2020\|EPI_ISL_660035\|20 | ···················· | ···················· | ····················· | ···················· | ···················· | ···················· |
| S | 6876 | hCoV-19/England/QEUH-B2F6C2/2020\|EPI_ISL_660036\|20 | ···················· | ···················· | ····················· | ···················· | ···················· | ···················· |
| S | 6877 | hCoV-19/England/QEUH-B303D6/2020\|EPI_ISL_660037\|20 | ···················· | ···················· | ····················· | ···················· | ···················· | ···················· |
| S | 6878 | hCoV-19/England/QEUH-B2CDDF/2020\|EPI_ISL_660038\|20 | ···················· | ···················· | ····················· | ···················· | ···················· | ···················· |
| S | 6879 | hCoV-19/England/QEUH-B2E863/2020\|EPI_ISL_660039\|20 | ···················· | ···················· | ····················· | ···················· | ···················· | ···················· |
| S | 6880 | hCoV-19/England/QEUH-B2D460/2020\|EPI_ISL_660040\|20 | ···················· | ···················· | ····················· | ···················· | ···················· | ···················· |
| S | 6881 | hCoV-19/England/QEUH-B2D855/2020\|EPI_ISL_660041\|20 | ···················· | ···················· | ····················· | ···················· | ···················· | ···················· |
| S | 6882 | hCoV-19/England/QEUH-B2D451/2020\|EPI_ISL_660043\|20 | ···················· | ·A·················· | ····················· | ···················· | ···················· | ··A················· |
| S | 6883 | hCoV-19/England/QEUH-B2FCB1/2020\|EPI_ISL_660044\|20 | ···················· | ·A·················· | ····················· | ···················· | ···················· | ···················· |
| S | 6884 | hCoV-19/England/QEUH-B2CD48/2020\|EPI_ISL_660045\|20 | ···················· | ···················· | ····················· | ···················· | ···················· | ···················· |
| S | 6885 | hCoV-19/England/QEUH-B2D2C0/2020\|EPI_ISL_660046\|20 | ···················· | ···················· | ····················· | ···················· | ···················· | ···················· |
| S | 6886 | hCoV-19/England/QEUH-B30B47/2020\|EPI_ISL_660047\|20 | ···················· | ···················· | ····················· | ···················· | ···················· | ···················· |
| S | 6887 | hCoV-19/England/QEUH-B3037C/2020\|EPI_ISL_660048\|20 | ···················· | ···················· | ····················· | ···················· | ···················· | ···················· |
| S | 6888 | hCoV-19/England/QEUH-B2D66A/2020\|EPI_ISL_660049\|20 | ···················· | ···················· | ····················· | ···················· | ···················· | ···················· |
| S | 6889 | hCoV-19/England/QEUH-B2FD90/2020\|EPI_ISL_660050\|20 | ···················· | ···················· | ····················· | ···················· | ···················· | ···················· |
| S | 6890 | hCoV-19/England/QEUH-B2CD93/2020\|EPI_ISL_660051\|20 | ···················· | ···················· | ····················· | ···················· | ···················· | ···················· |
| S | 6891 | hCoV-19/England/QEUH-B2F8EA/2020\|EPI_ISL_660052\|20 | ···················· | ···················· | ····················· | ···················· | ···················· | ···················· |
| S | 6892 | hCoV-19/England/QEUH-B2D57C/2020\|EPI_ISL_660053\|20 | ···················· | ·A·················· | ····················· | ···················· | ···················· | ···················· |
| S | 6893 | hCoV-19/England/QEUH-B2D512/2020\|EPI_ISL_660054\|20 | ···················· | ···················· | ····················· | ···················· | ······T············· | ···················· |
| S | 6894 | hCoV-19/Scotland/QEUH-B2CC78/2020\|EPI_ISL_660056\|2 | ···················· | ···················· | ····················· | ···················· | ···················· | ···················· |
| S | 6895 | hCoV-19/England/QEUH-B2D8FB/2020\|EPI_ISL_660057\|20 | ···················· | ···················· | ····················· | ···················· | ···················· | ···················· |
| S | 6896 | hCoV-19/England/QEUH-B30C08/2020\|EPI_ISL_660058\|20 | ···················· | ···················· | ····················· | ···················· | ···················· | ···················· |
| S | 6897 | hCoV-19/England/QEUH-B2F8BD/2020\|EPI_ISL_660059\|20 | ···················· | ···················· | ····················· | ···················· | ···················· | ···················· |
| S | 6898 | hCoV-19/England/QEUH-B2D442/2020\|EPI_ISL_660060\|20 | ···················· | ···················· | ····················· | ···················· | ···················· | ···················· |
| S | 6899 | hCoV-19/England/QEUH-B3043D/2020\|EPI_ISL_660061\|20 | ···················· | ···················· | ····················· | ···················· | ···················· | ···················· |
| S | 6900 | hCoV-19/England/QEUH-B2DFF6/2020\|EPI_ISL_660062\|20 | ···················· | ···················· | ····················· | ···················· | ···················· | ···················· |
| S | 6901 | hCoV-19/England/QEUH-B2CECD/2020\|EPI_ISL_660064\|20 | ···················· | ···················· | ····················· | ···················· | ···················· | ···················· |
| S | 6902 | hCoV-19/England/QEUH-B2EF9B/2020\|EPI_ISL_660065\|20 | ···················· | ·A·················· | ····················· | ···················· | ···················· | ···················· |
| S | 6903 | hCoV-19/Ecuador/ZZL-3/2020\|EPI_ISL_660069\|2020-07- | ···················· | ···················· | ····················· | ···················· | ···················· | ···················· |
| S | 6904 | hCoV-19/Ecuador/ZZL-4/2020\|EPI_ISL_660070\|2020-11- | ···················· | ···················· | ····················· | ···················· | ···················· | ···················· |
| S | 6905 | hCoV-19/USA/MD-MDH-0370/2020\|EPI_ISL_660071\|2020-1 | ···················· | ·A·················· | ····················· | ···················· | ···················· | ···················· |
| S | 6906 | hCoV-19/USA/MD-MDH-0371/2020\|EPI_ISL_660072\|2020-1 | ···················· | ···················· | ····················· | ···················· | ···················· | ···················· |
| S | 6907 | hCoV-19/USA/MD-MDH-0372/2020\|EPI_ISL_660073\|2020-1 | ···················· | ·A·················· | ····················· | ···················· | ···················· | ···················· |
| S | 6908 | hCoV-19/USA/MD-MDH-0373/2020\|EPI_ISL_660074\|2020-1 | ···················· | ···················· | ····················· | ···················· | ···················· | ···················· |
| S | 6909 | hCoV-19/USA/MD-MDH-0374/2020\|EPI_ISL_660075\|2020-1 | ···················· | ···················· | ····················· | ···················· | ···················· | ···················· |
| S | 6910 | hCoV-19/USA/MD-MDH-0375/2020\|EPI_ISL_660076\|2020-1 | ···················· | ···A················ | ····················· | ···················· | ···················· | ···················· |
| S | 6911 | hCoV-19/USA/MD-MDH-0376/2020\|EPI_ISL_660077\|2020-1 | ···················· | ···················· | ····················· | ···················· | ···················· | ···················· |
| S | 6912 | hCoV-19/USA/MD-MDH-0380/2020\|EPI_ISL_660078\|2020-1 | ···················· | ···················· | ···········T········· | ···················· | ···················· | ···················· |
| S | 6913 | hCoV-19/England/MILK-B0212E/2020\|EPI_ISL_649550\|20 | ···················· | ·A·················· | ····················· | ···················· | ···················· | ··A················· |

| | | | | | | | | |
|---|---|---|---|---|---|---|---|---|
| S | 6914 hCoV-19/England/MILK-B027A4/2020\|EPI_ISL_649551\|20 | ···················· | ···················· | ···················· | ···················· | ···················· | ···················· | |
| S | 6915 hCoV-19/England/MILK-B0321B/2020\|EPI_ISL_649552\|20 | ···················· | ···················· | ···················· | ···················· | ···················· | ···················· | |
| S | 6916 hCoV-19/England/MILK-B02D57/2020\|EPI_ISL_649553\|20 | ···················· | ···················· | ···················· | ···················· | ···················· | ···················· | |
| S | 6917 hCoV-19/England/MILK-B031E1/2020\|EPI_ISL_649554\|20 | ···················· | ···················· | ···················· | ···················· | ···················· | ···················· | |
| S | 6918 hCoV-19/England/MILK-B024E9/2020\|EPI_ISL_649555\|20 | ···················· | ···················· | ···················· | ···················· | ···················· | ···················· | |
| S | 6919 hCoV-19/Scotland/QEUH-A8BC0F/2020\|EPI_ISL_649556\|2 | ···················· | ···················· | ···················· | ···················· | ···················· | ···················· | |
| S | 6920 hCoV-19/Scotland/QEUH-A8B2A3/2020\|EPI_ISL_649557\|2 | ···················· | ···················· | ···················· | ···················· | ···················· | ···················· | |
| S | 6921 hCoV-19/England/QEUH-A8C855/2020\|EPI_ISL_649558\|20 | ···················· | ·R·················· | ···················· | ···················· | ···················· | ···················· | |
| S | 6922 hCoV-19/Scotland/QEUH-A8BEFA/2020\|EPI_ISL_649563\|2 | ···················· | ···················· | ···················· | ···················· | ···················· | ···················· | |
| S | 6923 hCoV-19/Scotland/QEUH-A8C12D/2020\|EPI_ISL_649564\|2 | ···················· | ···················· | ···················· | ···················· | ···················· | ···················· | |
| S | 6924 hCoV-19/Scotland/QEUH-B1059A/2020\|EPI_ISL_649600\|2 | ···················· | ···················· | ···················· | ···················· | ···················· | ···················· | |
| S | 6925 hCoV-19/Scotland/QEUH-B12914/2020\|EPI_ISL_649601\|2 | ···················· | ···················· | ···················· | ···················· | ···················· | ···················· | |
| S | 6926 hCoV-19/Scotland/QEUH-B13254/2020\|EPI_ISL_649602\|2 | ···················· | ···················· | ···················· | ···················· | ···················· | ···················· | |
| S | 6927 hCoV-19/Scotland/QEUH-B1362B/2020\|EPI_ISL_649603\|2 | ···················· | ···················· | ···················· | ···················· | ···················· | ···················· | |
| S | 6928 hCoV-19/Scotland/QEUH-B13315/2020\|EPI_ISL_649605\|2 | ···················· | ···················· | ···················· | ···················· | ···················· | ···················· | |
| S | 6929 hCoV-19/Scotland/CVR5602/2020\|EPI_ISL_651862\|2020- | ···················· | ···················· | ···················· | ···················· | ···················· | ···················· | |
| S | 6930 hCoV-19/Scotland/CVR5603/2020\|EPI_ISL_651863\|2020- | ···················· | ···················· | ···················· | ···················· | ···················· | ···················· | |
| S | 6931 hCoV-19/Scotland/CVR5605/2020\|EPI_ISL_651864\|2020- | ···················· | ···················· | ···················· | ···················· | ···················· | ···················· | |
| S | 6932 hCoV-19/Scotland/CVR5610/2020\|EPI_ISL_651865\|2020- | ···················· | ···················· | ···················· | ···················· | ···················· | ···················· | |
| S | 6933 hCoV-19/Scotland/CVR5611/2020\|EPI_ISL_651866\|2020- | ···················· | ···················· | ···················· | ···················· | ···················· | ···················· | |
| S | 6934 hCoV-19/Scotland/CVR5613/2020\|EPI_ISL_651867\|2020- | ···················· | ···················· | ···················· | ···················· | ···················· | ···················· | |
| S | 6935 hCoV-19/Scotland/CVR5615/2020\|EPI_ISL_651868\|2020- | ···················· | ···················· | ···················· | ···················· | ···················· | ···················· | |
| S | 6936 hCoV-19/Scotland/CVR5619/2020\|EPI_ISL_651869\|2020- | ···················· | ···················· | ···················· | ···················· | ···················· | ···················· | |
| S | 6937 hCoV-19/Scotland/CVR5620/2020\|EPI_ISL_651870\|2020- | ···················· | ···················· | ···················· | ···················· | ···················· | ···················· | |
| S | 6938 hCoV-19/Scotland/CVR5621/2020\|EPI_ISL_651871\|2020- | ···················· | ···················· | ···················· | ···················· | ···················· | ···················· | |
| S | 6939 hCoV-19/Scotland/CVR5622/2020\|EPI_ISL_651872\|2020- | ···················· | ···················· | ···················· | ···················· | ···················· | ···················· | |
| S | 6940 hCoV-19/Scotland/CVR5623/2020\|EPI_ISL_651873\|2020- | ···················· | ···················· | ···················· | ···················· | ···················· | ···················· | |
| S | 6941 hCoV-19/Scotland/CVR5626/2020\|EPI_ISL_651875\|2020- | ···················· | ···················· | ···················· | ···················· | ···················· | ···················· | |
| S | 6942 hCoV-19/Scotland/CVR5627/2020\|EPI_ISL_651876\|2020- | ···················· | ···················· | ···················· | ···················· | ···················· | ···················· | |
| S | 6943 hCoV-19/Scotland/CVR5631/2020\|EPI_ISL_651880\|2020- | ···················· | ···················· | ···················· | ···················· | ···················· | ···················· | |
| S | 6944 hCoV-19/Scotland/CVR5632/2020\|EPI_ISL_651881\|2020- | ···················· | ···················· | ···················· | ···················· | ···················· | ···················· | |
| S | 6945 hCoV-19/Scotland/CVR5633/2020\|EPI_ISL_651882\|2020- | ···················· | ···················· | ···················· | ···················· | ···················· | ···················· | |
| S | 6946 hCoV-19/Scotland/CVR5634/2020\|EPI_ISL_651883\|2020- | ···················· | ···················· | ···················· | ···················· | ···················· | ···················· | |
| S | 6947 hCoV-19/Scotland/CVR5676/2020\|EPI_ISL_651900\|2020- | ···················· | ···················· | ···················· | ···················· | ···················· | ···················· | |
| S | 6948 hCoV-19/Scotland/CVR5677/2020\|EPI_ISL_651901\|2020- | ···················· | ···················· | ···················· | ···················· | ···················· | ···················· | |
| S | 6949 hCoV-19/Scotland/CVR5679/2020\|EPI_ISL_651902\|2020- | ···················· | ···················· | ···················· | ···················· | ···················· | ···················· | |
| S | 6950 hCoV-19/Scotland/CVR5680/2020\|EPI_ISL_651903\|2020- | ···················· | ···················· | ···················· | ···················· | ···················· | ···················· | |
| S | 6951 hCoV-19/Scotland/CVR5681/2020\|EPI_ISL_651904\|2020- | ···················· | ···················· | ···················· | ···················· | ···················· | ···················· | |
| S | 6952 hCoV-19/Scotland/CVR5682/2020\|EPI_ISL_651905\|2020- | ···················· | ···················· | ···················· | ···················· | ···················· | ···················· | |
| S | 6953 hCoV-19/Scotland/CVR5683/2020\|EPI_ISL_651906\|2020- | ···················· | ···················· | ···················· | ···················· | ···················· | ···················· | |
| S | 6954 hCoV-19/Scotland/CVR5684/2020\|EPI_ISL_651907\|2020- | ···················· | ···················· | ···················· | ···················· | ···················· | ···················· | |
| S | 6955 hCoV-19/Scotland/CVR5685/2020\|EPI_ISL_651908\|2020- | ···················· | ···················· | ···················· | ···················· | ···················· | ···················· | |
| S | 6956 hCoV-19/Scotland/CVR5687/2020\|EPI_ISL_651909\|2020- | ···················· | ···················· | ···················· | ···················· | ···················· | ···················· | |
| S | 6957 hCoV-19/Scotland/CVR5688/2020\|EPI_ISL_651910\|2020- | ···················· | ···················· | ···················· | ···················· | ···················· | ···················· | |
| S | 6958 hCoV-19/Scotland/CVR5690/2020\|EPI_ISL_651911\|2020- | ···················· | ···················· | ···················· | ···················· | ···················· | ···················· | |
| S | 6959 hCoV-19/Scotland/CVR5691/2020\|EPI_ISL_651912\|2020- | ···················· | ···················· | ···················· | ···················· | ···················· | ···················· | |
| S | 6960 hCoV-19/Scotland/CVR5692/2020\|EPI_ISL_651913\|2020- | ···················· | ···················· | ···················· | ···················· | ···················· | ···················· | |
| S | 6961 hCoV-19/Scotland/CVR5693/2020\|EPI_ISL_651914\|2020- | ···················· | ···················· | ···················· | ···················· | ···················· | ···················· | |
| S | 6962 hCoV-19/Scotland/CVR5694/2020\|EPI_ISL_651915\|2020- | ···················· | ···················· | ···················· | ···················· | ···················· | ···················· | |
| S | 6963 hCoV-19/Scotland/CVR5695/2020\|EPI_ISL_651916\|2020- | ···················· | ···················· | ···················· | ···················· | ···················· | ···················· | |
| S | 6964 hCoV-19/Scotland/CVR5696/2020\|EPI_ISL_651917\|2020- | ···················· | ···················· | ···················· | ···················· | ···················· | ···················· | |
| S | 6965 hCoV-19/Scotland/CVR5698/2020\|EPI_ISL_651918\|2020- | ···················· | ···················· | ···················· | ···················· | ···················· | ···················· | |
| S | 6966 hCoV-19/Scotland/CVR5699/2020\|EPI_ISL_651919\|2020- | ···················· | ···················· | ···················· | ···················· | ···················· | ···················· | |
| S | 6970 hCoV-19/Spain/MD-IBV-99008570/2020\|EPI_ISL_654110\| | ···················· | ···················· | ···················· | ···················· | ···················· | ···················· | |
| S | 6972 hCoV-19/Spain/MD-IBV-99008572/2020\|EPI_ISL_654112\| | ···················· | ···················· | ···················· | ···················· | ···················· | ···················· | |
| S | 6974 hCoV-19/Spain/MD-IBV-99008574/2020\|EPI_ISL_654114\| | ···················· | ···················· | ···················· | ···················· | ···················· | ···················· | |
| S | 6977 hCoV-19/Spain/MD-IBV-99008577/2020\|EPI_ISL_654117\| | ···················· | ···················· | ···················· | ···················· | ···················· | ···················· | |
| S | 6981 hCoV-19/Spain/MD-IBV-99008581/2020\|EPI_ISL_654121\| | ···················· | ···················· | ···················· | ···················· | ·NN················· | ···················· | |
| S | 6982 hCoV-19/Spain/MD-IBV-99008582/2020\|EPI_ISL_654122\| | ···················· | ···················· | ···················· | ···················· | ···················· | ···················· | |
| S | 6984 hCoV-19/Spain/MD-IBV-99008585/2020\|EPI_ISL_654125\| | ···················· | ···················· | ···················· | ···················· | ···················· | ···················· | |
| S | 6987 hCoV-19/Spain/MD-IBV-99008588/2020\|EPI_ISL_654128\| | ···················· | ···················· | ···················· | ···················· | ···················· | ···················· | |
| S | 6988 hCoV-19/Spain/MD-IBV-99008589/2020\|EPI_ISL_654129\| | ···················· | ···················· | ···················· | ···················· | ···················· | ···················· | |
| S | 6989 hCoV-19/Spain/MD-IBV-99008590/2020\|EPI_ISL_654130\| | ···················· | ···················· | ···················· | ···················· | ···················· | ···················· | |
| S | 6992 hCoV-19/Spain/MD-IBV-99008593/2020\|EPI_ISL_654133\| | ···················· | ···················· | ···················· | ···················· | ···················· | ···················· | |
| S | 6993 hCoV-19/Spain/MD-IBV-99008594/2020\|EPI_ISL_654134\| | ···················· | ···················· | ···················· | ···················· | ·NNN················ | ···················· | 1 |
| S | 6995 hCoV-19/Spain/MD-IBV-99008597/2020\|EPI_ISL_654137\| | ···················· | ···················· | ···················· | ···················· | ···················· | ···················· | |
| S | 6996 hCoV-19/Spain/MD-IBV-99008599/2020\|EPI_ISL_654138\| | ···················· | ···················· | ···················· | ···················· | ···················· | ···················· | |
| S | 6999 hCoV-19/Spain/MD-IBV-99008791/2020\|EPI_ISL_654142\| | ···················· | ···················· | ···················· | ···················· | ···················· | ···················· | |
| S | 7000 hCoV-19/Spain/MD-IBV-99008792/2020\|EPI_ISL_654143\| | ···················· | ···················· | ···················· | ···················· | ···················· | ···················· | |
| S | 7003 hCoV-19/Spain/MD-IBV-99008795/2020\|EPI_ISL_654146\| | ···················· | ···················· | ···················· | ···················· | ···················· | ···················· | |
| S | 7005 hCoV-19/Spain/MD-IBV-99008797/2020\|EPI_ISL_654148\| | ···················· | ···················· | ···················· | ···················· | ···················· | ···················· | |
| S | 7007 hCoV-19/Spain/MD-IBV-99008799/2020\|EPI_ISL_654150\| | ···················· | ···················· | ···················· | ···················· | ···················· | ···················· | |
| S | 7008 hCoV-19/Spain/MD-IBV-99008800/2020\|EPI_ISL_654151\| | ···················· | ···················· | ···················· | ···················· | ···················· | ···················· | |
| S | 7009 hCoV-19/Spain/MD-IBV-99008801/2020\|EPI_ISL_654152\| | ···················· | ···················· | ···················· | ···················· | ···················· | ···················· | |
| S | 7010 hCoV-19/Spain/MD-IBV-99008802/2020\|EPI_ISL_654153\| | ···················· | ···················· | ···················· | ···················· | ···················· | ···················· | |
| S | 7011 hCoV-19/Spain/MD-IBV-99008803/2020\|EPI_ISL_654154\| | ···················· | ···················· | ···················· | ···················· | ···················· | ···················· | |
| S | 7012 hCoV-19/Spain/MD-IBV-99008804/2020\|EPI_ISL_654155\| | ···················· | ···················· | ···················· | ···················· | ···················· | ···················· | |
| S | 7013 hCoV-19/Spain/MD-IBV-99008805/2020\|EPI_ISL_654156\| | ···················· | ···················· | ···················· | ···················· | ···················· | ···················· | |
| S | 7019 hCoV-19/Spain/MD-IBV-99008607/2020\|EPI_ISL_654186\| | ···················· | ···················· | ···················· | ···················· | ···················· | ···················· | |
| S | 7024 hCoV-19/Spain/MD-IBV-99008610/2020\|EPI_ISL_654191\| | ···················· | ···················· | ···················· | ···················· | ···················· | ···················· | |

| | | | | | | | | | |
|---|---|---|---|---|---|---|---|---|---|
| S | 7026 | hCoV-19/Spain/MD-IBV-99008614/2020\|EPI_ISL_654193\| | ···················· | ···················· | ····················· | ···················· | ···················· | ···················· | |
| S | 7027 | hCoV-19/Spain/MD-IBV-99008615/2020\|EPI_ISL_654194\| | ···················· | ···················· | ····················· | ···················· | ···················· | ···················· | |
| S | 7028 | hCoV-19/Spain/MD-IBV-99008617/2020\|EPI_ISL_654195\| | ···················· | ···················· | ····················· | ···················· | ···················· | ···················· | |
| S | 7030 | hCoV-19/Spain/MD-IBV-99008621/2020\|EPI_ISL_654197\| | ···················· | ···················· | ····················· | ···················· | ···················· | ···················· | |
| S | 7032 | hCoV-19/Spain/MD-IBV-99008624/2020\|EPI_ISL_654199\| | ···················· | ···················· | ····················· | ···················· | ···················· | ···················· | |
| S | 7033 | hCoV-19/England/ALDP-B62BE2/2020\|EPI_ISL_655400\|20 | ···················· | ···················· | ····················· | ···················· | ···················· | ···················· | |
| S | 7034 | hCoV-19/England/ALDP-B62D28/2020\|EPI_ISL_655401\|20 | ···················· | ···················· | ····················· | ···················· | ···················· | ···················· | |
| S | 7035 | hCoV-19/England/ALDP-B62DCE/2020\|EPI_ISL_655404\|20 | ···················· | ···················· | ····················· | ···················· | ···················· | ···················· | |
| S | 7036 | hCoV-19/England/ALDP-B5E6E8/2020\|EPI_ISL_655405\|20 | ···················· | ···················· | ····················· | ···················· | ···················· | ···················· | |
| S | 7037 | hCoV-19/England/ALDP-B5E74F/2020\|EPI_ISL_655408\|20 | ···················· | ···················· | ····················· | ···················· | ···················· | ···················· | |
| S | 7038 | hCoV-19/England/ALDP-B5B1BD/2020\|EPI_ISL_655409\|20 | ···················· | ···················· | ····················· | ···················· | ···················· | ···················· | |
| S | 7039 | hCoV-19/England/ALDP-B5BA0D/2020\|EPI_ISL_655410\|20 | ···················· | ···················· | ····················· | ···················· | ···················· | ···················· | |
| S | 7040 | hCoV-19/England/ALDP-B5B047/2020\|EPI_ISL_655411\|20 | ···················· | ···················· | ····················· | ···················· | ···················· | ···················· | |
| S | 7041 | hCoV-19/England/ALDP-B5BDE6/2020\|EPI_ISL_655412\|20 | ···················· | ···················· | ····················· | ···················· | ···················· | ···················· | |
| S | 7042 | hCoV-19/England/ALDP-B5BD13/2020\|EPI_ISL_655416\|20 | ···················· | ···················· | ····················· | ···················· | ···················· | ···················· | |
| S | 7043 | hCoV-19/England/ALDP-B5B065/2020\|EPI_ISL_655417\|20 | ···················· | ···················· | ····················· | ···················· | ···················· | ···················· | |
| S | 7044 | hCoV-19/England/ALDP-B62E61/2020\|EPI_ISL_655418\|20 | ···················· | ···················· | ····················· | ···················· | ···················· | ···················· | |
| S | 7045 | hCoV-19/England/ALDP-B62B79/2020\|EPI_ISL_655419\|20 | ··················T· | ···················· | ····················· | ···················· | ···················· | ···················· | 1 |
| S | 7046 | hCoV-19/England/ALDP-B5B609/2020\|EPI_ISL_655420\|20 | ···················· | ···················· | ····················· | ···················· | ···················· | ···················· | |
| S | 7047 | hCoV-19/England/ALDP-B5E6CA/2020\|EPI_ISL_655421\|20 | ···················· | ···················· | ····················· | ···················· | ···················· | ···················· | |
| S | 7048 | hCoV-19/England/ALDP-B62B88/2020\|EPI_ISL_655422\|20 | ···················· | ···················· | ····················· | ···················· | ···················· | ··A················· | |
| S | 7049 | hCoV-19/England/ALDP-B62C2B/2020\|EPI_ISL_655423\|20 | ···················· | ···················· | ····················· | ···················· | ···················· | ···················· | |
| S | 7050 | hCoV-19/England/ALDP-B5B6CD/2020\|EPI_ISL_655424\|20 | ···················· | ···················· | ····················· | ···················· | ···················· | ···················· | |
| S | 7051 | hCoV-19/England/ALDP-B5E68E/2020\|EPI_ISL_655425\|20 | ···················· | ···················· | ····················· | ···················· | ···················· | ···················· | |
| S | 7052 | hCoV-19/England/ALDP-B62D0A/2020\|EPI_ISL_655426\|20 | ···················· | ···················· | ····················· | ···················· | ···················· | ···················· | |
| S | 7053 | hCoV-19/England/ALDP-B5B29C/2020\|EPI_ISL_655427\|20 | ···················· | ···················· | ····················· | ···················· | ···················· | ···················· | |
| S | 7054 | hCoV-19/England/ALDP-B62D91/2020\|EPI_ISL_655438\|20 | ···················· | ···················· | ····················· | ···················· | ···················· | ···················· | |
| S | 7055 | hCoV-19/England/ALDP-B63C57/2020\|EPI_ISL_655441\|20 | ···················· | ···················· | ····················· | ···················· | ···················· | ···················· | |
| S | 7056 | hCoV-19/England/ALDP-B5BA67/2020\|EPI_ISL_655445\|20 | ···················· | ···················· | ····················· | ···················· | ···················· | ···················· | |
| S | 7057 | hCoV-19/England/ALDP-B5B399/2020\|EPI_ISL_655446\|20 | ···················· | ···················· | ····················· | ···················· | ···················· | ···················· | |
| S | 7058 | hCoV-19/England/ALDP-B5BDC8/2020\|EPI_ISL_655448\|20 | ···················· | ···················· | ····················· | ···················· | ···················· | ···················· | |
| S | 7059 | hCoV-19/England/ALDP-B5B0CF/2020\|EPI_ISL_655449\|20 | ···················· | ···················· | ····················· | ···················· | ···················· | ···················· | |
| S | 7060 | hCoV-19/England/ALDP-B5B2BA/2020\|EPI_ISL_655451\|20 | ···················· | ···················· | ····················· | ···················· | ···················· | ···················· | |
| S | 7061 | hCoV-19/England/ALDP-B6304C/2020\|EPI_ISL_655454\|20 | ···················· | ···················· | ····················· | ···················· | ···················· | ···················· | |
| S | 7062 | hCoV-19/England/ALDP-B5B45A/2020\|EPI_ISL_655456\|20 | ···················· | ···················· | ····················· | ···················· | ···················· | ···················· | |
| S | 7063 | hCoV-19/England/ALDP-B63BB4/2020\|EPI_ISL_655457\|20 | ···················· | ···················· | ····················· | ···················· | ···················· | ···················· | |
| S | 7064 | hCoV-19/England/ALDP-B62C58/2020\|EPI_ISL_655459\|20 | ···················· | ···················· | ····················· | ···················· | ···················· | ···················· | |
| S | 7065 | hCoV-19/England/ALDP-B5B9A6/2020\|EPI_ISL_655460\|20 | ···················· | ···················· | ····················· | ···················· | ···················· | ···················· | |
| S | 7066 | hCoV-19/England/ALDP-B5AF78/2020\|EPI_ISL_655462\|20 | ···················· | ···················· | ····················· | ···················· | ···················· | ···················· | |
| S | 7067 | hCoV-19/England/ALDP-B5AF2D/2020\|EPI_ISL_655463\|20 | ···················· | ···················· | ····················· | ···················· | ···················· | ···················· | |
| S | 7068 | hCoV-19/England/ALDP-B5B627/2020\|EPI_ISL_655464\|20 | ···················· | ···················· | ····················· | ···················· | ···················· | ···················· | |
| S | 7069 | hCoV-19/England/ALDP-B5B0A1/2020\|EPI_ISL_655465\|20 | ···················· | ···················· | ····················· | ···················· | ···················· | ···················· | |
| S | 7070 | hCoV-19/England/ALDP-B5B557/2020\|EPI_ISL_655466\|20 | ···················· | ···················· | ····················· | ···················· | ···················· | ···················· | |
| S | 7071 | hCoV-19/England/ALDP-B5B86D/2020\|EPI_ISL_655467\|20 | ···················· | ···················· | ····················· | ···················· | ···················· | ···················· | |
| S | 7072 | hCoV-19/England/ALDP-B5BBCE/2020\|EPI_ISL_655468\|20 | ···················· | ···················· | ····················· | ···················· | ···················· | ···················· | |
| S | 7073 | hCoV-19/England/ALDP-B5E78B/2020\|EPI_ISL_655469\|20 | ···················· | ···················· | ····················· | ···················· | ···················· | ···················· | |
| S | 7074 | hCoV-19/England/ALDP-B62CEF/2020\|EPI_ISL_655470\|20 | ···················· | ···················· | ····················· | ···················· | ···················· | ···················· | |
| S | 7075 | hCoV-19/England/ALDP-B5B92E/2020\|EPI_ISL_655471\|20 | ···················· | ···················· | ····················· | ···················· | ···················· | ···················· | |
| S | 7076 | hCoV-19/England/ALDP-B6303D/2020\|EPI_ISL_655472\|20 | ···················· | ···················· | ····················· | ···················· | ···················· | ···················· | |
| S | 7077 | hCoV-19/England/ALDP-B5B126/2020\|EPI_ISL_655473\|20 | ···················· | ···················· | ····················· | ···················· | ···················· | ···················· | |
| S | 7078 | hCoV-19/England/ALDP-B5B584/2020\|EPI_ISL_655474\|20 | ···················· | ···················· | ····················· | ···················· | ···················· | ···················· | |
| S | 7079 | hCoV-19/England/ALDP-B5BD04/2020\|EPI_ISL_655475\|20 | ···················· | ···················· | ····················· | ···················· | ···················· | ···················· | |
| S | 7080 | hCoV-19/England/ALDP-B62F8C/2020\|EPI_ISL_655476\|20 | ···················· | ···················· | ····················· | ···················· | ···················· | ···················· | |
| S | 7081 | hCoV-19/England/ALDP-B5E79A/2020\|EPI_ISL_655477\|20 | ···················· | ···················· | ····················· | ···················· | ···················· | ···················· | |
| S | 7082 | hCoV-19/England/ALDP-B62F31/2020\|EPI_ISL_655478\|20 | ···················· | ···················· | ····················· | ···················· | ···················· | ···················· | |
| S | 7083 | hCoV-19/England/ALDP-B5E703/2020\|EPI_ISL_655479\|20 | ···················· | ···················· | ····················· | ···················· | ···················· | ···················· | |
| S | 7084 | hCoV-19/England/ALDP-B5BDAA/2020\|EPI_ISL_655481\|20 | ···················· | ···················· | ····················· | ···················· | ···················· | ···················· | |
| S | 7085 | hCoV-19/England/ALDP-B5B28D/2020\|EPI_ISL_655482\|20 | ···················· | ···················· | ····················· | ···················· | ···················· | ···················· | |
| S | 7086 | hCoV-19/England/ALDP-B5B979/2020\|EPI_ISL_655484\|20 | ···················· | ···················· | ····················· | ···················· | ···················· | ···················· | |
| S | 7087 | hCoV-19/England/ALDP-B62B97/2020\|EPI_ISL_655485\|20 | ···················· | ···················· | ····················· | ···················· | ···················· | ···················· | |
| S | 7088 | hCoV-19/England/ALDP-B62B3D/2020\|EPI_ISL_655486\|20 | ··················T· | ···················· | ····················· | ···················· | ···················· | ···················· | 1 |
| S | 7089 | hCoV-19/England/ALDP-B5B42D/2020\|EPI_ISL_655488\|20 | ···················· | ···················· | ····················· | ···················· | ···················· | ···················· | |
| S | 7090 | hCoV-19/England/ALDP-B5B320/2020\|EPI_ISL_655489\|20 | ···················· | ···················· | ····················· | ···················· | ···················· | ···················· | |
| S | 7091 | hCoV-19/England/ALDP-B62DDD/2020\|EPI_ISL_655490\|20 | ···················· | ···················· | ····················· | ···················· | ···················· | ···················· | |
| S | 7092 | hCoV-19/England/ALDP-B5B223/2020\|EPI_ISL_655491\|20 | ···················· | ···················· | ····················· | ···················· | ···················· | ···················· | |
| S | 7093 | hCoV-19/England/ALDP-B62D46/2020\|EPI_ISL_655492\|20 | ···················· | ···················· | ····················· | ···················· | ···················· | ···················· | |
| S | 7094 | hCoV-19/England/ALDP-B5BD8C/2020\|EPI_ISL_655493\|20 | ···················· | ···················· | ····················· | ···················· | ···················· | ···················· | |
| S | 7095 | hCoV-19/England/ALDP-B62FE6/2020\|EPI_ISL_655494\|20 | ···················· | ···················· | ····················· | ···················· | ···················· | ···················· | |
| S | 7096 | hCoV-19/England/ALDP-B5BC07/2020\|EPI_ISL_655496\|20 | ···················· | ···················· | ····················· | ···················· | ···················· | ···················· | |
| S | 7097 | hCoV-19/England/ALDP-B5B3F3/2020\|EPI_ISL_655497\|20 | ···················· | ···················· | ····················· | ···················· | ···················· | ···················· | |
| S | 7098 | hCoV-19/England/ALDP-B5B9F1/2020\|EPI_ISL_655499\|20 | ···················· | ···················· | ····················· | ···················· | ···················· | ···················· | |
| S | 7099 | hCoV-19/England/CAMC-B543AF/2020\|EPI_ISL_656701\|20 | ···················· | ·A·················· | ····················· | ···················· | ···················· | ··A················· | |
| S | 7100 | hCoV-19/England/CAMC-B53531/2020\|EPI_ISL_656702\|20 | ···················· | ···················· | ····················· | ···················· | ···················· | ···················· | |
| S | 7101 | hCoV-19/England/CAMC-B52462/2020\|EPI_ISL_656704\|20 | ···················· | ···················· | ····················· | ···················· | ·····T·············· | ···················· | |
| S | 7102 | hCoV-19/England/MILK-B5671A/2020\|EPI_ISL_656705\|20 | ···················· | ···················· | ····················· | ···················· | ···················· | ···················· | |
| S | 7103 | hCoV-19/England/CAMC-B54169/2020\|EPI_ISL_656706\|20 | ···················· | ···················· | ····················· | ···················· | ···················· | ···················· | |
| S | 7104 | hCoV-19/England/CAMC-B54354/2020\|EPI_ISL_656708\|20 | ···················· | ···················· | ····················· | ···················· | ···················· | ···················· | |
| S | 7105 | hCoV-19/England/CAMC-B540A8/2020\|EPI_ISL_656710\|20 | ···················· | ···················· | ····················· | ···················· | ···················· | ···················· | |
| S | 7106 | hCoV-19/England/CAMC-B5273C/2020\|EPI_ISL_656711\|20 | ···················· | ···················· | ····················· | ···················· | ···················· | ···················· | |
| S | 7107 | hCoV-19/England/CAMC-B53F8E/2020\|EPI_ISL_656712\|20 | ···················· | ···················· | ····················· | ···················· | ···················· | ···················· | |

| | | | | | | | |
|---|---|---|---|---|---|---|---|
| S | 7108 hCoV-19/England/CAMC-B53962/2020\|EPI_ISL_656713\|20 | ···················· | ···················· | ···················· | ···················· | ···················· | ···················· |
| S | 7109 hCoV-19/England/CAMC-B537B3/2020\|EPI_ISL_656714\|20 | ···················· | ···················· | ···················· | ···················· | ···················· | ···················· |
| S | 7110 hCoV-19/England/CAMC-B538B0/2020\|EPI_ISL_656715\|20 | ···················· | ···················· | ···················· | ···················· | ···················· | ···················· |
| S | 7111 hCoV-19/England/CAMC-B53926/2020\|EPI_ISL_656718\|20 | ···················· | ···················· | ···················· | ···················· | ···················· | ···················· |
| S | 7112 hCoV-19/England/CAMC-B526E4/2020\|EPI_ISL_656719\|20 | ···················· | ···················· | ···················· | ···················· | ···················· | ···················· |
| S | 7113 hCoV-19/England/CAMC-B53908/2020\|EPI_ISL_656720\|20 | ···················· | ···················· | ···················· | ···················· | ···················· | ···················· |
| S | 7114 hCoV-19/England/CAMC-B540C6/2020\|EPI_ISL_656721\|20 | ···················· | ···················· | ···················· | ···················· | ···················· | ···················· |
| S | 7115 hCoV-19/England/CAMC-B52286/2020\|EPI_ISL_656722\|20 | ···················· | ···················· | ···················· | ···················· | ···················· | ···················· |
| S | 7116 hCoV-19/England/CAMC-B5224A/2020\|EPI_ISL_656724\|20 | ···················· | ···················· | ···················· | ···················· | ···················· | ···················· |
| S | 7117 hCoV-19/England/CAMC-B53689/2020\|EPI_ISL_656725\|20 | ···················· | ···················· | ···················· | ···················· | ···················· | ···················· |
| S | 7118 hCoV-19/England/CAMC-B5359B/2020\|EPI_ISL_656726\|20 | ···················· | ···················· | ···················· | ···················· | ···················· | ···················· |
| S | 7119 hCoV-19/England/CAMC-B52426/2020\|EPI_ISL_656727\|20 | ···················· | ···················· | ···················· | ···················· | ···················· | ···················· |
| S | 7120 hCoV-19/England/MILK-B56A20/2020\|EPI_ISL_656728\|20 | ···················· | ···················· | ···················· | ···················· | ···················· | ···················· |
| S | 7121 hCoV-19/England/MILK-B56C93/2020\|EPI_ISL_656731\|20 | ···················· | ···················· | ···················· | ···················· | ···················· | ···················· |
| S | 7122 hCoV-19/England/MILK-B56950/2020\|EPI_ISL_656732\|20 | ···················· | ···················· | ···················· | ···················· | ···················· | ···················· |
| S | 7123 hCoV-19/England/MILK-B56B1E/2020\|EPI_ISL_656733\|20 | ···················· | ···················· | ···················· | ···················· | ···················· | ···················· |
| S | 7124 hCoV-19/England/CAMC-B535E6/2020\|EPI_ISL_656734\|20 | ···················· | ···················· | ···················· | ···················· | ···················· | ···················· |
| S | 7125 hCoV-19/England/CAMC-B5230B/2020\|EPI_ISL_656735\|20 | ···················· | ···················· | ···················· | ···················· | ···················· | ···················· |
| S | 7126 hCoV-19/England/CAMC-B522C2/2020\|EPI_ISL_656736\|20 | ···················· | ···················· | ···················· | ···················· | ···················· | ···················· |
| S | 7127 hCoV-19/England/CAMC-B5413C/2020\|EPI_ISL_656737\|20 | ···················· | ···················· | ···················· | ···················· | ···················· | ···················· |
| S | 7128 hCoV-19/England/CAMC-B53865/2020\|EPI_ISL_656739\|20 | ···················· | ···················· | ···················· | ···················· | ···················· | ···················· |
| S | 7129 hCoV-19/England/CAMC-B5266C/2020\|EPI_ISL_656740\|20 | ···················· | ···················· | ···················· | ···················· | ···················· | ···················· |
| S | 7130 hCoV-19/England/CAMC-B52259/2020\|EPI_ISL_656741\|20 | ···················· | ···················· | ···················· | ···················· | ···················· | ···················· |
| S | 7131 hCoV-19/England/CAMC-B536C5/2020\|EPI_ISL_656742\|20 | ···················· | ···················· | ···················· | ···················· | ···················· | ···················· |
| S | 7132 hCoV-19/England/CAMC-B526A8/2020\|EPI_ISL_656743\|20 | ···················· | ···················· | ···················· | ···················· | ···················· | ···················· |
| S | 7133 hCoV-19/England/MILK-B56C66/2020\|EPI_ISL_656744\|20 | ···················· | ···················· | ···················· | ···················· | ···················· | ···················· |
| S | 7134 hCoV-19/England/MILK-B56808/2020\|EPI_ISL_656745\|20 | ···················· | ···················· | ···················· | ···················· | ···················· | ···················· |
| S | 7135 hCoV-19/England/MILK-B568AE/2020\|EPI_ISL_656746\|20 | ···················· | ···················· | ···················· | ···················· | ···················· | ···················· |
| S | 7136 hCoV-19/England/CAMC-B523DE/2020\|EPI_ISL_656747\|20 | ···················· | ···················· | ···················· | ···················· | ···················· | ···················· |
| S | 7137 hCoV-19/England/CAMC-B544D9/2020\|EPI_ISL_656748\|20 | ···················· | ···················· | ···················· | ···················· | ···················· | ···················· |
| S | 7138 hCoV-19/England/CAMC-B54433/2020\|EPI_ISL_656750\|20 | ···················· | ···················· | ···················· | ···················· | ···················· | ···················· |
| S | 7139 hCoV-19/England/CAMC-B54512/2020\|EPI_ISL_656751\|20 | ···················· | ···················· | ···················· | ···················· | ···················· | ···················· |
| S | 7140 hCoV-19/England/CAMC-B525F6/2020\|EPI_ISL_656752\|20 | ···················· | ···················· | ···················· | ···················· | ···················· | ···················· |
| S | 7141 hCoV-19/England/CAMC-B54521/2020\|EPI_ISL_656754\|20 | ···················· | ···················· | ···················· | ···················· | ···················· | ···················· |
| S | 7142 hCoV-19/England/CAMC-B5415A/2020\|EPI_ISL_656755\|20 | ···················· | ···················· | ···················· | ···················· | ···················· | ···················· |
| S | 7143 hCoV-19/England/CAMC-B54530/2020\|EPI_ISL_656756\|20 | ···················· | ···················· | ···················· | ···················· | ···················· | ···················· |
| S | 7144 hCoV-19/England/MILK-B56B78/2020\|EPI_ISL_656757\|20 | ···················· | ···················· | ···················· | ···················· | ···················· | ···················· |
| S | 7145 hCoV-19/England/CAMC-B52611/2020\|EPI_ISL_656758\|20 | ···················· | ···················· | ···················· | ···················· | ···················· | ···················· |
| S | 7146 hCoV-19/England/CAMC-B5408A/2020\|EPI_ISL_656768\|20 | ···················· | ···················· | ···················· | ···················· | ···················· | ···················· |
| S | 7147 hCoV-19/England/CAMC-B5258D/2020\|EPI_ISL_656769\|20 | ···················· | ···················· | ···················· | ···················· | ···················· | ···················· |
| S | 7148 hCoV-19/England/CAMC-B54284/2020\|EPI_ISL_656771\|20 | ···················· | ···················· | ···················· | ···················· | ···················· | ···················· |
| S | 7149 hCoV-19/England/CAMC-B535AA/2020\|EPI_ISL_656772\|20 | ···················· | ···················· | ···················· | ···················· | ·····T·············· | ···················· |
| S | 7150 hCoV-19/England/CAMC-B540F3/2020\|EPI_ISL_656773\|20 | ···················· | ···················· | ···················· | ···················· | ···················· | ···················· |
| S | 7151 hCoV-19/England/MILK-B56B0F/2020\|EPI_ISL_656774\|20 | ···················· | ···················· | ···················· | ···················· | ···················· | ···················· |
| S | 7152 hCoV-19/England/CAMC-B53461/2020\|EPI_ISL_656775\|20 | ···················· | ···················· | ···················· | ···················· | ·····T·············· | ···················· |
| S | 7153 hCoV-19/England/MILK-B56765/2020\|EPI_ISL_656777\|20 | ···················· | ···················· | ···················· | ···················· | ···················· | ···················· |
| S | 7154 hCoV-19/England/MILK-B56B4B/2020\|EPI_ISL_656778\|20 | ···················· | ···················· | ···················· | ···················· | ···················· | ···················· |
| S | 7155 hCoV-19/England/MILK-B56BC3/2020\|EPI_ISL_656779\|20 | ···················· | ···················· | ···················· | ···················· | ···················· | ···················· |
| S | 7156 hCoV-19/England/CAMC-B53FF7/2020\|EPI_ISL_656782\|20 | ···················· | ·A·················· | ···················· | ···················· | ···················· | ··A················· |
| S | 7157 hCoV-19/England/CAMC-B543FA/2020\|EPI_ISL_656784\|20 | ···················· | ···················· | ···················· | ···················· | ···················· | ···················· |
| S | 7158 hCoV-19/England/CAMC-B53698/2020\|EPI_ISL_656787\|20 | ···················· | ···················· | ···················· | ···················· | ···················· | ···················· |
| S | 7159 hCoV-19/England/MILK-B56B87/2020\|EPI_ISL_656788\|20 | ···················· | ···················· | ···················· | ···················· | ···················· | ···················· |
| S | 7160 hCoV-19/England/MILK-B56A4E/2020\|EPI_ISL_656789\|20 | ···················· | ···················· | ···················· | ···················· | ···················· | ···················· |
| S | 7161 hCoV-19/England/CAMC-B542FD/2020\|EPI_ISL_656790\|20 | ···················· | ···················· | ···················· | ···················· | ···················· | ···················· |
| S | 7162 hCoV-19/England/CAMC-B5373B/2020\|EPI_ISL_656792\|20 | ···················· | ···················· | ···················· | ···················· | ···················· | ···················· |
| S | 7163 hCoV-19/England/CAMC-B5456D/2020\|EPI_ISL_656793\|20 | ···················· | ···················· | ···················· | ···················· | ···················· | ···················· |
| S | 7164 hCoV-19/England/CAMC-B54099/2020\|EPI_ISL_656794\|20 | ···················· | ···················· | ···················· | ···················· | ···················· | ···················· |
| S | 7165 hCoV-19/England/CAMC-B5420C/2020\|EPI_ISL_656795\|20 | ···················· | ···················· | ···················· | ···················· | ···················· | ···················· |
| S | 7166 hCoV-19/England/CAMC-B5380B/2020\|EPI_ISL_656797\|20 | ···················· | ·A·······A·········· | ···················· | ···················· | ···················· | ···················· |
| S | 7167 hCoV-19/England/MILK-B56A5D/2020\|EPI_ISL_656799\|20 | ···················· | ···················· | ···················· | ···················· | ···················· | ···················· |
| S | 7168 hCoV-19/Serbia/Pancevo-009210204/2020\|EPI_ISL_6612 | ···················· | ···················· | ···················· | ···················· | ···················· | ···················· |
| S | 7169 hCoV-19/Scotland/QEUH-B13B4A/2020\|EPI_ISL_649500\|2 | ···················· | ···················· | ···················· | ···················· | ···················· | ···················· |
| S | 7170 hCoV-19/Scotland/QEUH-B12FD6/2020\|EPI_ISL_649501\|2 | ···················· | ···················· | ···················· | ···················· | ···················· | ···················· |
| S | 7171 hCoV-19/Scotland/QEUH-B0EFFB/2020\|EPI_ISL_649502\|2 | ···················· | ···················· | ···················· | ···················· | ···················· | ···················· |
| S | 7172 hCoV-19/Scotland/QEUH-B0F07F/2020\|EPI_ISL_649504\|2 | ···················· | ···················· | ···················· | ···················· | ···················· | ···················· |
| S | 7173 hCoV-19/Scotland/QEUH-B13F20/2020\|EPI_ISL_649505\|2 | ···················· | ···················· | ···················· | ···················· | ···················· | ···················· |
| S | 7174 hCoV-19/Scotland/QEUH-B1064C/2020\|EPI_ISL_649506\|2 | ···················· | ···················· | ···················· | ···················· | ···················· | ···················· |
| S | 7175 hCoV-19/Scotland/QEUH-B0EFDD/2020\|EPI_ISL_649507\|2 | ···················· | ···················· | ···················· | ···················· | ···················· | ···················· |
| S | 7176 hCoV-19/Scotland/QEUH-B1303C/2020\|EPI_ISL_649508\|2 | ···················· | ···················· | ···················· | ···················· | ···················· | ···················· |
| S | 7177 hCoV-19/Scotland/QEUH-B13C38/2020\|EPI_ISL_649509\|2 | ···················· | ···················· | ···················· | ···················· | ···················· | ···················· |
| S | 7178 hCoV-19/Scotland/QEUH-B12FA9/2020\|EPI_ISL_649510\|2 | ···················· | ···················· | ···················· | ···················· | ···················· | ···················· |
| S | 7179 hCoV-19/Scotland/QEUH-B0F08E/2020\|EPI_ISL_649511\|2 | ···················· | ···················· | ···················· | ···················· | ···················· | ···················· |
| S | 7180 hCoV-19/England/QEUH-B1352E/2020\|EPI_ISL_649512\|20 | ···················· | ···················· | ···················· | ···················· | ···················· | ···················· |
| S | 7181 hCoV-19/Scotland/QEUH-B106D3/2020\|EPI_ISL_649513\|2 | ···················· | ···················· | ···················· | ···················· | ···················· | ···················· |
| S | 7182 hCoV-19/Scotland/QEUH-B0F024/2020\|EPI_ISL_649514\|2 | ···················· | ···················· | ···················· | ···················· | ···················· | ···················· |
| S | 7183 hCoV-19/Scotland/QEUH-B1066A/2020\|EPI_ISL_649515\|2 | ···················· | ···················· | ···················· | ···················· | ···················· | ···················· |
| S | 7184 hCoV-19/Scotland/QEUH-B0F015/2020\|EPI_ISL_649516\|2 | ···················· | ···················· | ···················· | ···················· | ···················· | ···················· |
| S | 7185 hCoV-19/Scotland/QEUH-B0DAEC/2020\|EPI_ISL_649517\|2 | ···················· | ···················· | ···················· | ···················· | ···················· | ···················· |
| S | 7186 hCoV-19/Scotland/QEUH-B0DA64/2020\|EPI_ISL_649518\|2 | ···················· | ···················· | ···················· | ···················· | ···················· | ···················· |
| S | 7187 hCoV-19/Scotland/QEUH-B0F2A6/2020\|EPI_ISL_649519\|2 | ···················· | ···················· | ···················· | ···················· | ···················· | ···················· |

| | | | | | | | |
|---|---|---|---|---|---|---|---|
| S | 7188 hCoV-19/Scotland/QEUH-B0DB25/2020\|EPI_ISL_649520\|2 | ···················· | ···················· | ···················· | ···················· | ···················· | ···················· |
| S | 7189 hCoV-19/Scotland/QEUH-B0F288/2020\|EPI_ISL_649521\|2 | ···················· | ···················· | ···················· | ···················· | ···················· | ···················· |
| S | 7190 hCoV-19/Scotland/QEUH-B110A7/2020\|EPI_ISL_649522\|2 | ···················· | ···················· | ···················· | ···················· | ···················· | ···················· |
| S | 7191 hCoV-19/Scotland/QEUH-B0EC5E/2020\|EPI_ISL_649523\|2 | ···················· | ···················· | ···················· | ···················· | ···················· | ···················· |
| S | 7192 hCoV-19/Scotland/QEUH-B0DA91/2020\|EPI_ISL_649525\|2 | ···················· | ···················· | ···················· | ···················· | ···················· | ···················· |
| S | 7193 hCoV-19/Scotland/QEUH-B0F297/2020\|EPI_ISL_649526\|2 | ···················· | ···················· | ···················· | ···················· | ···················· | ···················· |
| S | 7194 hCoV-19/Scotland/QEUH-B0DD5C/2020\|EPI_ISL_649527\|2 | ···················· | ···················· | ···················· | ···················· | ···················· | ···················· |
| S | 7195 hCoV-19/Scotland/QEUH-B1105C/2020\|EPI_ISL_649528\|2 | ···················· | ···················· | ···················· | ···················· | ···················· | ···················· |
| S | 7196 hCoV-19/Scotland/QEUH-B11010/2020\|EPI_ISL_649529\|2 | ···················· | ···················· | ···················· | ···················· | ···················· | ···················· |
| S | 7197 hCoV-19/Scotland/QEUH-B0F2B5/2020\|EPI_ISL_649530\|2 | ···················· | ···················· | ···················· | ···················· | ···················· | ···················· |
| S | 7198 hCoV-19/Scotland/QEUH-B0DA46/2020\|EPI_ISL_649531\|2 | ···················· | ···················· | ···················· | ···················· | ···················· | ···················· |
| S | 7199 hCoV-19/Scotland/QEUH-B0DB07/2020\|EPI_ISL_649532\|2 | ···················· | ·····A·············· | ···················· | ···················· | ···················· | ···················· |
| S | 7200 hCoV-19/Scotland/QEUH-B0F279/2020\|EPI_ISL_649533\|2 | ···················· | ···················· | ···················· | ···················· | ···················· | ···················· |
| S | 7201 hCoV-19/Scotland/QEUH-B10FAB/2020\|EPI_ISL_649534\|2 | ···················· | ···················· | ···················· | ···················· | ···················· | ···················· |
| S | 7202 hCoV-19/Scotland/QEUH-B10F9C/2020\|EPI_ISL_649535\|2 | ···················· | ···················· | ···················· | ···················· | ···················· | ···················· |
| S | 7203 hCoV-19/Scotland/QEUH-B10FF6/2020\|EPI_ISL_649536\|2 | ···················· | ···················· | ···················· | ···················· | ···················· | ···················· |
| S | 7204 hCoV-19/Scotland/QEUH-B11001/2020\|EPI_ISL_649538\|2 | ···················· | ···················· | ···················· | ···················· | ···················· | ···················· |
| S | 7205 hCoV-19/Scotland/QEUH-B1102F/2020\|EPI_ISL_649539\|2 | ···················· | ···················· | ···················· | ···················· | ···················· | ···················· |
| S | 7206 hCoV-19/Scotland/QEUH-B0DCC8/2020\|EPI_ISL_649540\|2 | ···················· | ···················· | ···················· | ···················· | ···················· | ···················· |
| S | 7207 hCoV-19/Scotland/QEUH-B0E386/2020\|EPI_ISL_649541\|2 | ···················· | ···················· | ···················· | ···················· | ···················· | ···················· |
| S | 7208 hCoV-19/Scotland/QEUH-B0DA82/2020\|EPI_ISL_649542\|2 | ···················· | ···················· | ···················· | ···················· | ···················· | ···················· |
| S | 7209 hCoV-19/Scotland/QEUH-B0DABF/2020\|EPI_ISL_649544\|2 | ···················· | ···················· | ···················· | ···················· | ···················· | ···················· |
| S | 7210 hCoV-19/Scotland/QEUH-B10FD8/2020\|EPI_ISL_649545\|2 | ···················· | ···················· | ···················· | ···················· | ···················· | ···················· |
| S | 7211 hCoV-19/Scotland/QEUH-B10FC9/2020\|EPI_ISL_649546\|2 | ···················· | ···················· | ···················· | ···················· | ···················· | ···················· |
| S | 7212 hCoV-19/Scotland/QEUH-B1103E/2020\|EPI_ISL_649547\|2 | ···················· | ···················· | ···················· | ···················· | ···················· | ···················· |
| S | 7213 hCoV-19/England/MILK-B06342/2020\|EPI_ISL_649548\|20 | ···················· | ···················· | ···················· | ···················· | ·····T·············· | ···················· |
| S | 7214 hCoV-19/Scotland/MILK-B027D1/2020\|EPI_ISL_649549\|2 | ···················· | ···················· | ···················· | ···················· | ···················· | ···················· |
| S | 7215 hCoV-19/Scotland/QEUH-A8B2D0/2020\|EPI_ISL_649560\|2 | ···················· | ···················· | ···················· | ···················· | ···················· | ···················· |
| S | 7216 hCoV-19/Scotland/QEUH-A8B8A1/2020\|EPI_ISL_649561\|2 | ···················· | ···················· | ···················· | ···················· | ···················· | ··A················· |
| S | 7217 hCoV-19/Scotland/QEUH-A8B2C1/2020\|EPI_ISL_649562\|2 | ···················· | ···················· | ···················· | ···················· | ···················· | ···················· |
| S | 7218 hCoV-19/Scotland/QEUH-A8BDEE/2020\|EPI_ISL_649568\|2 | ···················· | ···················· | ···················· | ···················· | ···················· | ···················· |
| S | 7219 hCoV-19/Scotland/QEUH-B10AE6/2020\|EPI_ISL_649569\|2 | ···················· | ···················· | ···················· | ···················· | ···················· | ···················· |
| S | 7220 hCoV-19/Scotland/QEUH-B11405/2020\|EPI_ISL_649570\|2 | ···················· | ···················· | ···················· | ···················· | ···················· | ···················· |
| S | 7221 hCoV-19/Scotland/QEUH-B132AF/2020\|EPI_ISL_649571\|2 | ···················· | ···················· | ···················· | ···················· | ···················· | ···················· |
| S | 7222 hCoV-19/Scotland/QEUH-B129BA/2020\|EPI_ISL_649572\|2 | ···················· | ···················· | ···················· | ···················· | ···················· | ···················· |
| S | 7223 hCoV-19/Scotland/QEUH-B118DC/2020\|EPI_ISL_649573\|2 | ···················· | ···················· | ···················· | ···················· | ···················· | ···················· |
| S | 7224 hCoV-19/Scotland/QEUH-B13263/2020\|EPI_ISL_649574\|2 | ···················· | ···················· | ···················· | ···················· | ···················· | ···················· |
| S | 7225 hCoV-19/Scotland/QEUH-B12738/2020\|EPI_ISL_649575\|2 | ···················· | ···················· | ···················· | ···················· | ···················· | ···················· |
| S | 7226 hCoV-19/Scotland/QEUH-B11DDD/2020\|EPI_ISL_649576\|2 | ···················· | ···················· | ···················· | ···················· | ···················· | ···················· |
| S | 7227 hCoV-19/Scotland/QEUH-B121D0/2020\|EPI_ISL_649577\|2 | ···················· | ···················· | ···················· | ···················· | ···················· | ···················· |
| S | 7228 hCoV-19/Scotland/QEUH-B10AC8/2020\|EPI_ISL_649578\|2 | ···················· | ···················· | ···················· | ···················· | ···················· | ···················· |
| S | 7229 hCoV-19/Scotland/QEUH-B132BE/2020\|EPI_ISL_649579\|2 | ···················· | ···················· | ···················· | ···················· | ·····T·············· | ···················· |
| S | 7230 hCoV-19/Scotland/QEUH-B128EA/2020\|EPI_ISL_649581\|2 | ···················· | ···················· | ···················· | ···················· | ···················· | ···················· |
| S | 7231 hCoV-19/Scotland/QEUH-B10AD7/2020\|EPI_ISL_649582\|2 | ···················· | ···················· | ···················· | ···················· | ···················· | ···················· |
| S | 7232 hCoV-19/Scotland/QEUH-B105B8/2020\|EPI_ISL_649583\|2 | ···················· | ···················· | ···················· | ···················· | ···················· | ···················· |
| S | 7233 hCoV-19/Scotland/QEUH-B10AAA/2020\|EPI_ISL_649584\|2 | ···················· | ···················· | ···················· | ···················· | ···················· | ···················· |
| S | 7234 hCoV-19/Scotland/QEUH-B127CF/2020\|EPI_ISL_649585\|2 | ···················· | ···················· | ···················· | ···················· | ···················· | ···················· |
| S | 7235 hCoV-19/Scotland/QEUH-B13290/2020\|EPI_ISL_649586\|2 | ···················· | ···················· | ···················· | ···················· | ···················· | ···················· |
| S | 7236 hCoV-19/Scotland/QEUH-B10AF5/2020\|EPI_ISL_649587\|2 | ···················· | ···················· | ···················· | ···················· | ···················· | ···················· |
| S | 7237 hCoV-19/Scotland/QEUH-B132EB/2020\|EPI_ISL_649588\|2 | ···················· | ···················· | ···················· | ···················· | ···················· | ···················· |
| S | 7238 hCoV-19/Scotland/QEUH-B132FA/2020\|EPI_ISL_649589\|2 | ···················· | ···················· | ···················· | ···················· | ···················· | ···················· |
| S | 7239 hCoV-19/Scotland/QEUH-B13281/2020\|EPI_ISL_649590\|2 | ···················· | ···················· | ···················· | ···················· | ···················· | ···················· |
| S | 7240 hCoV-19/Scotland/QEUH-B11E07/2020\|EPI_ISL_649592\|2 | ···················· | ···················· | ···················· | ···················· | ···················· | ···················· |
| S | 7241 hCoV-19/Scotland/QEUH-B135E2/2020\|EPI_ISL_649593\|2 | ···················· | ·A·················· | ···················· | ···················· | ···················· | ···················· |
| S | 7242 hCoV-19/Scotland/QEUH-B13324/2020\|EPI_ISL_649594\|2 | ···················· | ···················· | ···················· | ···················· | ···················· | ···················· |
| S | 7243 hCoV-19/Scotland/QEUH-B1055E/2020\|EPI_ISL_649595\|2 | ···················· | ···················· | ···················· | ···················· | ···················· | ···················· |
| S | 7244 hCoV-19/Scotland/QEUH-B10B10/2020\|EPI_ISL_649596\|2 | ···················· | ···················· | ···················· | ···················· | ···················· | ···················· |
| S | 7245 hCoV-19/Scotland/QEUH-B10B01/2020\|EPI_ISL_649598\|2 | ···················· | ···················· | ···················· | ···················· | ···················· | ···················· |
| S | 7246 hCoV-19/Scotland/QEUH-B13272/2020\|EPI_ISL_649599\|2 | ···················· | ···················· | ···················· | ···················· | ···················· | ···················· |
| S | 7247 hCoV-19/Scotland/CVR5401/2020\|EPI_ISL_651800\|2020- | ···················· | ···················· | ···················· | ···················· | ···················· | ···················· |
| S | 7248 hCoV-19/Scotland/CVR5402/2020\|EPI_ISL_651801\|2020- | ···················· | ···················· | ···················· | ···················· | ···················· | ···················· |
| S | 7249 hCoV-19/Scotland/CVR5403/2020\|EPI_ISL_651802\|2020- | ···················· | ···················· | ···················· | ···················· | ···················· | ···················· |
| S | 7250 hCoV-19/Scotland/CVR5405/2020\|EPI_ISL_651803\|2020- | ···················· | ···················· | ···················· | ···················· | ···················· | ···················· |
| S | 7251 hCoV-19/Scotland/CVR5408/2020\|EPI_ISL_651804\|2020- | ···················· | ···················· | ···················· | ···················· | ···················· | ···················· |
| S | 7252 hCoV-19/Scotland/CVR5409/2020\|EPI_ISL_651805\|2020- | ···················· | ···················· | ···················· | ···················· | ···················· | ···················· |
| S | 7253 hCoV-19/Scotland/CVR5411/2020\|EPI_ISL_651806\|2020- | ···················· | ···················· | ···················· | ···················· | ···················· | ···················· |
| S | 7254 hCoV-19/Scotland/CVR5412/2020\|EPI_ISL_651807\|2020- | ···················· | ···················· | ···················· | ···················· | ···················· | ···················· |
| S | 7255 hCoV-19/Scotland/CVR5413/2020\|EPI_ISL_651808\|2020- | ···················· | ···················· | ···················· | ···················· | ···················· | ···················· |
| S | 7256 hCoV-19/Scotland/CVR5414/2020\|EPI_ISL_651809\|2020- | ···················· | ···················· | ···················· | ···················· | ···················· | ···················· |
| S | 7257 hCoV-19/Scotland/CVR5416/2020\|EPI_ISL_651810\|2020- | ···················· | ···················· | ···················· | ···················· | ···················· | ···················· |
| S | 7258 hCoV-19/Scotland/CVR5474/2020\|EPI_ISL_651811\|2020- | ···················· | ···················· | ···················· | ···················· | ···················· | ···················· |
| S | 7259 hCoV-19/Scotland/CVR5478/2020\|EPI_ISL_651812\|2020- | ···················· | ···················· | ···················· | ···················· | ···················· | ···················· |
| S | 7260 hCoV-19/Scotland/CVR5485/2020\|EPI_ISL_651813\|2020- | ···················· | ···················· | ···················· | ···················· | ···················· | ···················· |
| S | 7261 hCoV-19/Scotland/CVR5489/2020\|EPI_ISL_651814\|2020- | ···················· | ···················· | ···················· | ···················· | ···················· | ···················· |
| S | 7262 hCoV-19/Scotland/CVR5490/2020\|EPI_ISL_651815\|2020- | ···················· | ···················· | ···················· | ···················· | ···················· | ···················· |
| S | 7263 hCoV-19/Scotland/CVR5491/2020\|EPI_ISL_651816\|2020- | ···················· | ···················· | ···················· | ···················· | ···················· | ···················· |
| S | 7264 hCoV-19/Scotland/CVR5493/2020\|EPI_ISL_651817\|2020- | ···················· | ···················· | ···················· | ···················· | ···················· | ···················· |
| S | 7265 hCoV-19/Scotland/CVR5494/2020\|EPI_ISL_651818\|2020- | ···················· | ···················· | ···················· | ···················· | ···················· | ···················· |
| S | 7266 hCoV-19/Scotland/CVR5495/2020\|EPI_ISL_651819\|2020- | ···················· | ···················· | ···················· | ···················· | ···················· | ···················· |
| S | 7267 hCoV-19/Scotland/CVR5496/2020\|EPI_ISL_651820\|2020- | ···················· | ···················· | ···················· | ···················· | ···················· | ···················· |

| | | | | | | | |
|---|---|---|---|---|---|---|---|
| S | 7268 hCoV-19/Scotland/CVR5512/2020\|EPI_ISL_651821\|2020- | ···················· | ···················· | ····················· | ···················· | ···················· | ···················· |
| S | 7269 hCoV-19/Scotland/CVR5513/2020\|EPI_ISL_651822\|2020- | ···················· | ···················· | ····················· | ···················· | ···················· | ···················· |
| S | 7270 hCoV-19/Scotland/CVR5514/2020\|EPI_ISL_651823\|2020- | ···················· | ···················· | ····················· | ···················· | ···················· | ···················· |
| S | 7271 hCoV-19/Scotland/CVR5515/2020\|EPI_ISL_651824\|2020- | ···················· | ···················· | ····················· | ···················· | ···················· | ···················· |
| S | 7272 hCoV-19/Scotland/CVR5516/2020\|EPI_ISL_651825\|2020- | ···················· | ···················· | ····················· | ···················· | ···················· | ···················· |
| S | 7273 hCoV-19/Scotland/CVR5519/2020\|EPI_ISL_651826\|2020- | ···················· | ···················· | ····················· | ···················· | ···················· | ···················· |
| S | 7274 hCoV-19/Scotland/CVR5521/2020\|EPI_ISL_651827\|2020- | ···················· | ···················· | ····················· | ···················· | ···················· | ···················· |
| S | 7275 hCoV-19/Scotland/CVR5524/2020\|EPI_ISL_651828\|2020- | ···················· | ···················· | ····················· | ···················· | ···················· | ···················· |
| S | 7276 hCoV-19/Scotland/CVR5528/2020\|EPI_ISL_651829\|2020- | ···················· | ···················· | ····················· | ···················· | ···················· | ···················· |
| S | 7277 hCoV-19/Scotland/CVR5537/2020\|EPI_ISL_651830\|2020- | ···················· | ···················· | ····················· | ···················· | ···················· | ···················· |
| S | 7278 hCoV-19/Scotland/CVR5541/2020\|EPI_ISL_651831\|2020- | ···················· | ···················· | ····················· | ···················· | ···················· | ···················· |
| S | 7279 hCoV-19/Scotland/CVR5545/2020\|EPI_ISL_651832\|2020- | ···················· | ···················· | ····················· | ···················· | ···················· | ···················· |
| S | 7280 hCoV-19/Scotland/CVR5546/2020\|EPI_ISL_651833\|2020- | ···················· | ···················· | ····················· | ···················· | ···················· | ···················· |
| S | 7281 hCoV-19/Scotland/CVR5548/2020\|EPI_ISL_651834\|2020- | ···················· | ···················· | ····················· | ···················· | ···················· | ···················· |
| S | 7282 hCoV-19/Scotland/CVR5551/2020\|EPI_ISL_651835\|2020- | ···················· | ···················· | ····················· | ···················· | ···················· | ···················· |
| S | 7283 hCoV-19/Scotland/CVR5557/2020\|EPI_ISL_651836\|2020- | ···················· | ···················· | ····················· | ···················· | ···················· | ···················· |
| S | 7284 hCoV-19/Scotland/CVR5560/2020\|EPI_ISL_651837\|2020- | ···················· | ···················· | ····················· | ···················· | ···················· | ···················· |
| S | 7285 hCoV-19/Scotland/CVR5561/2020\|EPI_ISL_651838\|2020- | ···················· | ···················· | ····················· | ···················· | ···················· | ···················· |
| S | 7286 hCoV-19/Scotland/CVR5562/2020\|EPI_ISL_651839\|2020- | ···················· | ···················· | ····················· | ···················· | ···················· | ···················· |
| S | 7287 hCoV-19/Scotland/CVR5563/2020\|EPI_ISL_651840\|2020- | ···················· | ···················· | ····················· | ···················· | ···················· | ···················· |
| S | 7288 hCoV-19/Scotland/CVR5564/2020\|EPI_ISL_651841\|2020- | ···················· | ···················· | ····················· | ···················· | ···················· | ···················· |
| S | 7289 hCoV-19/Scotland/CVR5567/2020\|EPI_ISL_651842\|2020- | ···················· | ···················· | ····················· | ···················· | ···················· | ···················· |
| S | 7290 hCoV-19/Scotland/CVR5568/2020\|EPI_ISL_651843\|2020- | ···················· | ···················· | ····················· | ···················· | ···················· | ···················· |
| S | 7291 hCoV-19/Scotland/CVR5569/2020\|EPI_ISL_651844\|2020- | ···················· | ···················· | ····················· | ···················· | ···················· | ···················· |
| S | 7292 hCoV-19/Scotland/CVR5571/2020\|EPI_ISL_651845\|2020- | ···················· | ···················· | ····················· | ···················· | ···················· | ···················· |
| S | 7293 hCoV-19/Scotland/CVR5572/2020\|EPI_ISL_651846\|2020- | ···················· | ···················· | ····················· | ···················· | ···················· | ···················· |
| S | 7294 hCoV-19/Scotland/CVR5574/2020\|EPI_ISL_651847\|2020- | ···················· | ···················· | ····················· | ···················· | ···················· | ···················· |
| S | 7295 hCoV-19/Scotland/CVR5576/2020\|EPI_ISL_651848\|2020- | ···················· | ···················· | ····················· | ···················· | ···················· | ···················· |
| S | 7296 hCoV-19/Scotland/CVR5579/2020\|EPI_ISL_651849\|2020- | ···················· | ···················· | ····················· | ···················· | ···················· | ···················· |
| S | 7297 hCoV-19/Scotland/CVR5584/2020\|EPI_ISL_651850\|2020- | ···················· | ···················· | ····················· | ···················· | ···················· | ···················· |
| S | 7298 hCoV-19/Scotland/CVR5586/2020\|EPI_ISL_651851\|2020- | ···················· | ···················· | ····················· | ···················· | ···················· | ···················· |
| S | 7299 hCoV-19/Scotland/CVR5587/2020\|EPI_ISL_651852\|2020- | ···················· | ···················· | ····················· | ···················· | ···················· | ···················· |
| S | 7300 hCoV-19/Scotland/CVR5588/2020\|EPI_ISL_651853\|2020- | ···················· | ···················· | ····················· | ···················· | ···················· | ···················· |
| S | 7301 hCoV-19/Scotland/CVR5589/2020\|EPI_ISL_651854\|2020- | ···················· | ···················· | ····················· | ···················· | ···················· | ···················· |
| S | 7302 hCoV-19/Scotland/CVR5591/2020\|EPI_ISL_651855\|2020- | ···················· | ···················· | ····················· | ···················· | ···················· | ···················· |
| S | 7303 hCoV-19/Scotland/CVR5593/2020\|EPI_ISL_651856\|2020- | ···················· | ···················· | ····················· | ···················· | ···················· | ···················· |
| S | 7304 hCoV-19/Scotland/CVR5594/2020\|EPI_ISL_651857\|2020- | ···················· | ···················· | ····················· | ···················· | ···················· | ···················· |
| S | 7305 hCoV-19/Scotland/CVR5597/2020\|EPI_ISL_651858\|2020- | ···················· | ···················· | ····················· | ···················· | ···················· | ···················· |
| S | 7306 hCoV-19/Scotland/CVR5599/2020\|EPI_ISL_651859\|2020- | ···················· | ···················· | ····················· | ···················· | ···················· | ···················· |
| S | 7307 hCoV-19/Scotland/CVR5600/2020\|EPI_ISL_651860\|2020- | ···················· | ···················· | ····················· | ···················· | ···················· | ···················· |
| S | 7308 hCoV-19/Scotland/CVR5601/2020\|EPI_ISL_651861\|2020- | ···················· | ···················· | ····················· | ···················· | ···················· | ···················· |
| S | 7309 hCoV-19/Scotland/CVR5624/2020\|EPI_ISL_651874\|2020- | ···················· | ···················· | ····················· | ···················· | ···················· | ···················· |
| S | 7310 hCoV-19/Scotland/CVR5628/2020\|EPI_ISL_651877\|2020- | ···················· | ···················· | ····················· | ···················· | ···················· | ···················· |
| S | 7311 hCoV-19/Scotland/CVR5629/2020\|EPI_ISL_651878\|2020- | ···················· | ···················· | ····················· | ···················· | ···················· | ···················· |
| S | 7312 hCoV-19/Scotland/CVR5630/2020\|EPI_ISL_651879\|2020- | ···················· | ···················· | ····················· | ···················· | ···················· | ···················· |
| S | 7313 hCoV-19/Scotland/CVR5641/2020\|EPI_ISL_651884\|2020- | ···················· | ···················· | ····················· | ···················· | ···················· | ···················· |
| S | 7314 hCoV-19/Scotland/CVR5643/2020\|EPI_ISL_651885\|2020- | ···················· | ·A·················· | ····················· | ···················· | ···················· | ···················· |
| S | 7315 hCoV-19/Scotland/CVR5655/2020\|EPI_ISL_651886\|2020- | ···················· | ·A·················· | ····················· | ···················· | ···················· | ···················· |
| S | 7316 hCoV-19/Scotland/CVR5656/2020\|EPI_ISL_651887\|2020- | ···················· | ···················· | ····················· | ···················· | ···················· | ···················· |
| S | 7317 hCoV-19/Scotland/CVR5657/2020\|EPI_ISL_651888\|2020- | ···················· | ···················· | ····················· | ···················· | ···················· | ···················· |
| S | 7318 hCoV-19/Scotland/CVR5658/2020\|EPI_ISL_651889\|2020- | ···················· | ···················· | ····················· | ···················· | ···················· | ···················· |
| S | 7319 hCoV-19/Scotland/CVR5659/2020\|EPI_ISL_651890\|2020- | ···················· | ···················· | ····················· | ···················· | ···················· | ···················· |
| S | 7320 hCoV-19/Scotland/CVR5661/2020\|EPI_ISL_651891\|2020- | ···················· | ···················· | ····················· | ···················· | ···················· | ···················· |
| S | 7321 hCoV-19/Scotland/CVR5662/2020\|EPI_ISL_651892\|2020- | ···················· | ···················· | ····················· | ···················· | ···················· | ···················· |
| S | 7322 hCoV-19/Scotland/CVR5663/2020\|EPI_ISL_651893\|2020- | ···················· | ···················· | ····················· | ···················· | ···················· | ···················· |
| S | 7323 hCoV-19/Scotland/CVR5664/2020\|EPI_ISL_651894\|2020- | ···················· | ···················· | ····················· | ···················· | ···················· | ···················· |
| S | 7324 hCoV-19/Scotland/CVR5666/2020\|EPI_ISL_651895\|2020- | ···················· | ···················· | ····················· | ···················· | ···················· | ···················· |
| S | 7325 hCoV-19/Scotland/CVR5671/2020\|EPI_ISL_651896\|2020- | ···················· | ···················· | ····················· | ···················· | ···················· | ···················· |
| S | 7326 hCoV-19/Scotland/CVR5672/2020\|EPI_ISL_651897\|2020- | ···················· | ···················· | ····················· | ···················· | ···················· | ···················· |
| S | 7327 hCoV-19/Scotland/CVR5673/2020\|EPI_ISL_651898\|2020- | ···················· | ···················· | ····················· | ···················· | ···················· | ···················· |
| S | 7328 hCoV-19/Scotland/CVR5674/2020\|EPI_ISL_651899\|2020- | ···················· | ···················· | ····················· | ···················· | ···················· | ···················· |
| S | 7329 hCoV-19/Spain/MD-IBV-99008532/2020\|EPI_ISL_654080\| | ···················· | ···················· | ····················· | ···················· | ···················· | ···················· |
| S | 7335 hCoV-19/Spain/MD-IBV-99008540/2020\|EPI_ISL_654086\| | ···················· | ···················· | ····················· | ···················· | ···················· | ···················· |
| S | 7336 hCoV-19/Spain/MD-IBV-99008541/2020\|EPI_ISL_654087\| | ···················· | ···················· | ····················· | ···················· | ···················· | ···················· |
| S | 7337 hCoV-19/Spain/MD-IBV-99008542/2020\|EPI_ISL_654088\| | ···················· | ···················· | ····················· | ···················· | ···················· | ···················· |
| S | 7342 hCoV-19/Spain/MD-IBV-99008555/2020\|EPI_ISL_654096\| | ···················· | ···················· | ····················· | ···················· | ···················· | ···················· |
| S | 7343 hCoV-19/Spain/MD-IBV-99008556/2020\|EPI_ISL_654097\| | ···················· | ···················· | ····················· | ···················· | ···················· | ···················· |
| S | 7344 hCoV-19/Spain/MD-IBV-99008557/2020\|EPI_ISL_654098\| | ···················· | ···················· | ····················· | ···················· | ···················· | ···················· |
| S | 7345 hCoV-19/Spain/MD-IBV-99008558/2020\|EPI_ISL_654099\| | ···················· | ···················· | ····················· | ···················· | ···················· | ···················· |
| S | 7346 hCoV-19/England/ALDP-B5B40F/2020\|EPI_ISL_655305\|20 | ···················· | ···················· | ····················· | ···················· | ···················· | ···················· |
| S | 7347 hCoV-19/England/ALDP-B5B539/2020\|EPI_ISL_655306\|20 | ···················· | ···················· | ····················· | ···················· | ···················· | ···················· |
| S | 7348 hCoV-19/England/ALDP-B62BD3/2020\|EPI_ISL_655307\|20 | ···················· | ···················· | ····················· | ···················· | ···················· | ···················· |
| S | 7349 hCoV-19/England/ALDP-B63D18/2020\|EPI_ISL_655308\|20 | ···················· | ···················· | ····················· | ···················· | ···················· | ···················· |
| S | 7350 hCoV-19/England/ALDP-B5B7E8/2020\|EPI_ISL_655309\|20 | ···················· | ···················· | ····················· | ···················· | ···················· | ···················· |
| S | 7351 hCoV-19/England/ALDP-B62D64/2020\|EPI_ISL_655310\|20 | ···················· | ···················· | ····················· | ···················· | ···················· | ···················· |
| S | 7352 hCoV-19/England/ALDP-B62FD7/2020\|EPI_ISL_655311\|20 | ···················· | ···················· | ····················· | ···················· | ···················· | ···················· |
| S | 7353 hCoV-19/England/ALDP-B5B706/2020\|EPI_ISL_655312\|20 | ···················· | ···················· | ····················· | ···················· | ···················· | ···················· |
| S | 7354 hCoV-19/England/ALDP-B5BAB2/2020\|EPI_ISL_655314\|20 | ···················· | ···················· | ····················· | ···················· | ···················· | ···················· |
| S | 7355 hCoV-19/England/ALDP-B62CC1/2020\|EPI_ISL_655315\|20 | ···················· | ···················· | ····················· | ···················· | ···················· | ···················· |
| S | 7356 hCoV-19/England/ALDP-B5E75E/2020\|EPI_ISL_655320\|20 | ···················· | ···················· | ····················· | ···················· | ···················· | ···················· |

| | | | | | | | |
|---|---|---|---|---|---|---|---|
| S | 7357 hCoV-19/England/ALDP-B5B8A9/2020\|EPI_ISL_655321\|20 | ···················· | ···················· | ···················· | ···················· | ···················· | ···················· |
| S | 7358 hCoV-19/England/ALDP-B5BAEF/2020\|EPI_ISL_655322\|20 | ···················· | ·A·················· | ···················· | ···················· | ···················· | ···A················ |
| S | 7359 hCoV-19/England/ALDP-B62C85/2020\|EPI_ISL_655326\|20 | ···················· | ···················· | ···················· | ···················· | ···················· | ···················· |
| S | 7360 hCoV-19/England/ALDP-B62B00/2020\|EPI_ISL_656636\|20 | ···················· | ···················· | ···················· | ···················· | ···················· | ···················· |
| S | 7361 hCoV-19/England/ALDP-B624AB/2020\|EPI_ISL_656637\|20 | ···················· | ···················· | ···················· | ···················· | ······T············· | ···················· |
| S | 7362 hCoV-19/England/ALDP-B63273/2020\|EPI_ISL_656638\|20 | ···················· | ·A·················· | ···················· | ···················· | ···················· | ···················· |
| S | 7363 hCoV-19/England/ALDP-B636D1/2020\|EPI_ISL_656639\|20 | ···················· | ···················· | ···················· | ···················· | ···················· | ···················· |
| S | 7364 hCoV-19/England/ALDP-B639C9/2020\|EPI_ISL_656640\|20 | ···················· | ···················· | ···················· | ···················· | ···················· | ···················· |
| S | 7365 hCoV-19/England/ALDP-B63695/2020\|EPI_ISL_656641\|20 | ···················· | ···················· | ···················· | ···················· | ···················· | ···················· |
| S | 7366 hCoV-19/England/ALDP-B6339E/2020\|EPI_ISL_656642\|20 | ···················· | ···················· | ···················· | ···················· | ···················· | ···················· |
| S | 7367 hCoV-19/England/ALDP-B62A7C/2020\|EPI_ISL_656644\|20 | ···················· | ···················· | ···················· | ···················· | ···················· | ···················· |
| S | 7368 hCoV-19/England/ALDP-B63228/2020\|EPI_ISL_656645\|20 | ···················· | ···················· | ···················· | ···················· | ···················· | ···················· |
| S | 7370 hCoV-19/England/ALDP-B6211D/2020\|EPI_ISL_656650\|20 | ···················· | ···················· | ···················· | ···················· | ···················· | ···················· |
| S | 7371 hCoV-19/Wales/ALDP-B63862/2020\|EPI_ISL_656654\|2020 | ···················· | ·A·················· | ···················· | ···················· | ···················· | ···A················ |
| S | 7372 hCoV-19/England/ALDP-B622A1/2020\|EPI_ISL_656655\|20 | ···················· | ···················· | ···················· | ···················· | ···················· | ···················· |
| S | 7373 hCoV-19/England/ALDP-B624D8/2020\|EPI_ISL_656656\|20 | ···················· | ···················· | ···················· | ···················· | ···················· | ···················· |
| S | 7374 hCoV-19/England/ALDP-B63334/2020\|EPI_ISL_656657\|20 | ···················· | ···················· | ···················· | ···················· | ···················· | ···················· |
| S | 7375 hCoV-19/England/CAMC-B51F08/2020\|EPI_ISL_657967\|20 | ···················· | ···················· | ···················· | ···················· | ···················· | ···················· |
| S | 7376 hCoV-19/England/CAMC-B51366/2020\|EPI_ISL_657968\|20 | ···················· | ···················· | ···················· | ···················· | ···················· | ···················· |
| S | 7377 hCoV-19/England/MILK-B54C2C/2020\|EPI_ISL_657970\|20 | ···················· | ···················· | ···················· | ···················· | ···················· | ···················· |
| S | 7378 hCoV-19/England/MILK-B54E62/2020\|EPI_ISL_657971\|20 | ···················· | ···················· | ···················· | ···················· | ···················· | ···················· |
| S | 7379 hCoV-19/England/MILK-B54D83/2020\|EPI_ISL_657972\|20 | ···················· | ···················· | ···················· | ···················· | ···················· | ···················· |
| S | 7380 hCoV-19/England/MILK-B55344/2020\|EPI_ISL_657973\|20 | ···················· | ···················· | ···················· | ···················· | ···················· | ···················· |
| S | 7381 hCoV-19/England/CAMC-B52453/2020\|EPI_ISL_657974\|20 | ···················· | ·A·················· | ···················· | ···················· | ···················· | ···A················ |
| S | 7382 hCoV-19/England/MILK-B54E9F/2020\|EPI_ISL_657975\|20 | ···················· | ···················· | ···················· | ···················· | ···················· | ···················· |
| S | 7383 hCoV-19/England/MILK-B5553F/2020\|EPI_ISL_657977\|20 | ···················· | ···················· | ···················· | ···················· | ···················· | ···················· |
| S | 7384 hCoV-19/England/MILK-B54FBA/2020\|EPI_ISL_657980\|20 | ···················· | ···················· | ···················· | ···················· | ···················· | ···················· |
| S | 7385 hCoV-19/England/MILK-B54B5C/2020\|EPI_ISL_657981\|20 | ···················· | ···················· | ···················· | ···················· | ···················· | ···················· |
| S | 7386 hCoV-19/England/MILK-B5510E/2020\|EPI_ISL_657985\|20 | ···················· | ···················· | ···················· | ···················· | ···················· | ···················· |
| S | 7387 hCoV-19/England/CAMC-B51287/2020\|EPI_ISL_657987\|20 | ···················· | ···················· | ···················· | ···················· | ···················· | ···················· |
| S | 7388 hCoV-19/England/MILK-B54AC8/2020\|EPI_ISL_657988\|20 | ···················· | ···················· | ···················· | ···················· | ···················· | ···················· |
| S | 7389 hCoV-19/Belgium/ULG-10781/2020\|EPI_ISL_661218\|2020 | ···················· | ···················· | ···················· | ···················· | ···················· | ···················· |
| S | 7390 hCoV-19/Belgium/ULG-10783/2020\|EPI_ISL_661219\|2020 | ···················· | ···················· | ···················· | ···················· | ···················· | ···················· |
| S | 7391 hCoV-19/Belgium/ULG-10784/2020\|EPI_ISL_661220\|2020 | ···················· | ···················· | ···················· | ···················· | ···················· | ···················· |
| S | 7392 hCoV-19/Belgium/ULG-10785/2020\|EPI_ISL_661221\|2020 | ···················· | ·A·················· | ···················· | ···················· | ···················· | ···················· |
| S | 7393 hCoV-19/Belgium/ULG-10787/2020\|EPI_ISL_661222\|2020 | ···················· | ···················· | ···················· | ···················· | ···················· | ···················· |
| S | 7394 hCoV-19/Belgium/ULG-10788/2020\|EPI_ISL_661223\|2020 | ···················· | ····A··············· | ···················· | ···················· | ···················· | ···················· |
| S | 7395 hCoV-19/Belgium/ULG-10789/2020\|EPI_ISL_661224\|2020 | ···················· | ···················· | ···················· | ···················· | ···················· | ···················· |
| S | 7396 hCoV-19/Belgium/ULG-10790/2020\|EPI_ISL_661225\|2020 | ···················· | ···················· | ···················· | ···················· | ···················· | ···················· |
| S | 7397 hCoV-19/Belgium/ULG-10791/2020\|EPI_ISL_661226\|2020 | ···················· | ···················· | ···················· | ···················· | ···················· | ···················· |
| S | 7398 hCoV-19/Belgium/ULG-10793/2020\|EPI_ISL_661227\|2020 | ···················· | ···················· | ···················· | ···················· | ···C················ | ···················· |
| S | 7399 hCoV-19/Belgium/ULG-10794/2020\|EPI_ISL_661228\|2020 | ···················· | ···················· | ···················· | ···················· | ···················· | ···················· |
| S | 7400 hCoV-19/Belgium/ULG-10799/2020\|EPI_ISL_661229\|2020 | ···················· | ···················· | ···················· | ···················· | ···················· | ···················· |
| S | 7401 hCoV-19/Belgium/ULG-10800/2020\|EPI_ISL_661230\|2020 | ···················· | ···················· | ···················· | ···················· | ···················· | ···················· |
| S | 7402 hCoV-19/Belgium/ULG-10801/2020\|EPI_ISL_661231\|2020 | ···················· | ···················· | ···················· | ···················· | ···················· | ···················· |
| S | 7403 hCoV-19/Belgium/ULG-10803/2020\|EPI_ISL_661232\|2020 | ···················· | ···················· | ···················· | ···················· | ···················· | ···················· |
| S | 7404 hCoV-19/Belgium/ULG-10806/2020\|EPI_ISL_661233\|2020 | ···················· | ···················· | ···················· | ···················· | ···················· | ···················· |
| S | 7405 hCoV-19/Belgium/ULG-10807/2020\|EPI_ISL_661234\|2020 | ···················· | ···················· | ···················· | ···················· | ···················· | ···················· |
| S | 7406 hCoV-19/Belgium/ULG-10808/2020\|EPI_ISL_661235\|2020 | ···················· | ···················· | ···················· | ···················· | ···················· | ···················· |
| S | 7407 hCoV-19/Belgium/ULG-10811/2020\|EPI_ISL_661236\|2020 | ···················· | ···················· | ···················· | ···················· | ···················· | ···················· |
| S | 7408 hCoV-19/Belgium/ULG-10812/2020\|EPI_ISL_661237\|2020 | ···················· | ···················· | ···················· | ···················· | ···················· | ···················· |
| S | 7409 hCoV-19/Belgium/ULG-10813/2020\|EPI_ISL_661238\|2020 | ···················· | ···················· | ···················· | ···················· | ···················· | ···················· |
| S | 7410 hCoV-19/Belgium/ULG-10814/2020\|EPI_ISL_661239\|2020 | ···················· | ···················· | ···················· | ···················· | ···················· | ···················· |
| S | 7411 hCoV-19/Belgium/ULG-10816/2020\|EPI_ISL_661240\|2020 | ···················· | ···················· | ···················· | ···················· | ···················· | ···················· |
| S | 7412 hCoV-19/Belgium/ULG-10817/2020\|EPI_ISL_661241\|2020 | ···················· | ···················· | ···················· | ···················· | ···················· | ···················· |
| S | 7413 hCoV-19/Belgium/ULG-10819/2020\|EPI_ISL_661242\|2020 | ···················· | ···················· | ···················· | ···················· | ···················· | ···················· |
| S | 7414 hCoV-19/Belgium/ULG-10820/2020\|EPI_ISL_661243\|2020 | ···················· | ···················· | ···················· | ···················· | ···················· | ···················· |
| S | 7415 hCoV-19/Belgium/ULG-10821/2020\|EPI_ISL_661244\|2020 | ···················· | ···················· | ···················· | ···················· | ···················· | ···················· |
| S | 7416 hCoV-19/Belgium/ULG-10822/2020\|EPI_ISL_661245\|2020 | ···················· | ···················· | ···················· | ···················· | ···················· | ···················· |
| S | 7417 hCoV-19/Belgium/ULG-10825/2020\|EPI_ISL_661246\|2020 | ···················· | ···················· | ···················· | ···················· | ···················· | ···················· |
| S | 7418 hCoV-19/Belgium/ULG-10827/2020\|EPI_ISL_661247\|2020 | ···················· | ···················· | ···················· | ···················· | ···················· | ···················· |
| S | 7419 hCoV-19/Belgium/ULG-10830/2020\|EPI_ISL_661248\|2020 | ···················· | ···················· | ···················· | ···················· | ···················· | ···················· |
| S | 7420 hCoV-19/Belgium/ULG-10831/2020\|EPI_ISL_661249\|2020 | ···················· | ···················· | ···················· | ···················· | ···················· | ···················· |
| S | 7421 hCoV-19/Belgium/ULG-10833/2020\|EPI_ISL_661250\|2020 | ···················· | ···················· | ···················· | ···················· | ···················· | ···················· |
| S | 7422 hCoV-19/Belgium/ULG-10834/2020\|EPI_ISL_661251\|2020 | ···················· | ·········A·········· | ···················· | ···················· | ···················· | ···················· |
| S | 7423 hCoV-19/Sichuan/SC-MY-001/2020\|EPI_ISL_661271\|2020 | ···················· | ···················· | ···················· | ···················· | ···················· | ···················· |
| S | 7424 hCoV-19/Palestine/WB-AQU-P3/2020\|EPI_ISL_661272\|20 | ···················· | ···················· | ···················· | ···················· | ···················· | ···················· |
| S | 7425 hCoV-19/England/204481886/2020\|EPI_ISL_654000\|2020 | ···················· | ···················· | ···················· | ···················· | ···················· | ···················· |
| S | 7426 hCoV-19/England/204481927/2020\|EPI_ISL_654001\|2020 | ···················· | ···················· | ···················· | ···················· | ···················· | ···················· |
| S | 7427 hCoV-19/England/204481943/2020\|EPI_ISL_654002\|2020 | ···················· | ···················· | ···················· | ···················· | ···················· | ···················· |
| S | 7428 hCoV-19/England/204481944/2020\|EPI_ISL_654003\|2020 | ···················· | ···················· | ···················· | ···················· | ···················· | ···················· |
| S | 7429 hCoV-19/England/204481947/2020\|EPI_ISL_654004\|2020 | ···················· | ···················· | ···················· | ···················· | ···················· | ···················· |
| S | 7430 hCoV-19/England/204481949/2020\|EPI_ISL_654005\|2020 | ···················· | ···················· | ···················· | ···················· | ···················· | ···················· |
| S | 7431 hCoV-19/England/204481951/2020\|EPI_ISL_654006\|2020 | ···················· | ···················· | ···················· | ···················· | ···················· | ···················· |
| S | 7432 hCoV-19/Tunisia/58412/2020\|EPI_ISL_654018\|2020-11- | ···················· | ···················· | ···················· | ···················· | ···················· | ···················· |
| S | 7433 hCoV-19/Tunisia/55308/2020\|EPI_ISL_654019\|2020-11- | ···················· | ···················· | ···················· | ···················· | ···················· | ···················· |
| S | 7434 hCoV-19/Tunisia/53873/2020\|EPI_ISL_654020\|2020-10- | ···················· | ···················· | ···················· | ···················· | ···················· | ···················· |
| S | 7435 hCoV-19/Spain/MD-IBV-99008465/2020\|EPI_ISL_654053\| | ···················· | ···················· | ···················· | ···················· | ···················· | ···················· |
| S | 7440 hCoV-19/Spain/MD-IBV-99008475/2020\|EPI_ISL_654058\| | ···················· | ···················· | ···················· | ···················· | ·NNN················ | ···················· |
| S | 7441 hCoV-19/Spain/MD-IBV-99008476/2020\|EPI_ISL_654059\| | ···················· | ···················· | ···················· | ···················· | ···················· | ···················· |

S 7442 hCoV-19/Spain/MD-IBV-99008477/2020|EPI_ISL_654060| ···················· ···················· ····················· ···················· ···················· ····················
S 7444 hCoV-19/Spain/MD-IBV-99008480/2020|EPI_ISL_654062| ···················· ···················· ····················· ···················· ···················· ····················
S 7449 hCoV-19/Spain/MD-IBV-99008497/2020|EPI_ISL_654067| ···················· ···················· ····················· ···················· ···················· ····················
S 7454 hCoV-19/Spain/MD-IBV-99008507/2020|EPI_ISL_654072| ···················· ···················· ····················· ···················· ···················· ····················
S 7458 hCoV-19/Spain/MD-IBV-99008519/2020|EPI_ISL_654076| ···················· ···················· ····················· ···················· ···················· ····················
S 7459 hCoV-19/Spain/MD-IBV-99008521/2020|EPI_ISL_654077| ···················· ···················· ····················· ···················· ···················· ····················
S 7461 hCoV-19/Spain/MD-IBV-99008529/2020|EPI_ISL_654079| ···················· ···················· ····················· ···················· ···················· ····················
S 7462 hCoV-19/England/ALDP-B62CD0/2020|EPI_ISL_655300|20 ···················· ···················· ····················· ···················· ···················· ····················
S 7463 hCoV-19/England/ALDP-B62F04/2020|EPI_ISL_655301|20 ···················· ···················· ····················· ···················· ···A················ ····················
S 7464 hCoV-19/England/ALDP-B63DAF/2020|EPI_ISL_655302|20 ···················· ···················· ····················· ···················· ···················· ····················
S 7465 hCoV-19/England/ALDP-B5B78E/2020|EPI_ISL_655317|20 ···················· ···················· ····················· ···················· ···················· ····················
S 7466 hCoV-19/England/ALDP-B5B135/2020|EPI_ISL_655318|20 ···················· ···················· ····················· ···················· ···················· ····················
S 7467 hCoV-19/England/ALDP-B5AFF0/2020|EPI_ISL_655319|20 ···················· ···················· ····················· ···················· ···················· ····················
S 7468 hCoV-19/England/ALDP-B5B636/2020|EPI_ISL_655323|20 ···················· ···················· ····················· ···················· ···················· ····················
S 7469 hCoV-19/England/ALDP-B5B1CC/2020|EPI_ISL_655324|20 ···················· ···················· ····················· ···················· ···················· ····················
S 7470 hCoV-19/England/ALDP-B5B51B/2020|EPI_ISL_655325|20 ···················· ···················· ····················· ···················· ···················· ····················
S 7471 hCoV-19/England/ALDP-B5AEB7/2020|EPI_ISL_655328|20 ···················· ···················· ····················· ···················· ···················· ····················
S 7472 hCoV-19/England/ALDP-B5B79D/2020|EPI_ISL_655329|20 ···················· ···················· ····················· ···················· ···················· ····················
S 7473 hCoV-19/England/ALDP-B5E660/2020|EPI_ISL_655330|20 ···················· ···················· ····················· ···················· ···················· ····················
S 7474 hCoV-19/England/ALDP-B5BA3A/2020|EPI_ISL_655331|20 ···················· ···················· ····················· ···················· ···················· ····················
S 7475 hCoV-19/England/ALDP-B62B5B/2020|EPI_ISL_655333|20 ···················· ···················· ····················· ···················· ···················· ····················
S 7476 hCoV-19/England/ALDP-B5B654/2020|EPI_ISL_655334|20 ···················· ···················· ····················· ···················· ···················· ····················
S 7477 hCoV-19/England/ALDP-B5BA76/2020|EPI_ISL_655336|20 ···················· ···················· ····················· ···················· ···················· ····················
S 7478 hCoV-19/England/ALDP-B5BBA0/2020|EPI_ISL_655337|20 ···················· ···················· ····················· ···················· ···················· ····················
S 7479 hCoV-19/England/ALDP-B5B27E/2020|EPI_ISL_655340|20 ···················· ·A·················· ····················· ···················· ···················· ····················
S 7480 hCoV-19/England/ALDP-B5B0ED/2020|EPI_ISL_655344|20 ···················· ···················· ····················· ···················· ···················· ····················
S 7481 hCoV-19/England/ALDP-B5BB37/2020|EPI_ISL_655349|20 ···················· ·A·················· ····················· ···················· ···················· ···A················
S 7482 hCoV-19/England/ALDP-B5B4B4/2020|EPI_ISL_655350|20 ···················· ···················· ····················· ···················· ···················· ····················
S 7483 hCoV-19/England/ALDP-B5BB28/2020|EPI_ISL_655352|20 ···················· ···················· ····················· ···················· ···················· ····················
S 7484 hCoV-19/England/ALDP-B5B94C/2020|EPI_ISL_655353|20 ···················· ···················· ····················· ···················· ···················· ····················
S 7485 hCoV-19/England/ALDP-B5AED5/2020|EPI_ISL_655354|20 ···················· ···················· ····················· ···················· ···················· ····················
S 7486 hCoV-19/England/ALDP-B62AD6/2020|EPI_ISL_655355|20 ···················· ···················· ····················· ···················· ···················· ····················
S 7487 hCoV-19/England/ALDP-B5E76D/2020|EPI_ISL_655357|20 ···················· ···················· ····················· ···················· ···················· ····················
S 7488 hCoV-19/England/ALDP-B5B724/2020|EPI_ISL_655359|20 ···················· ···················· ····················· ···················· ···················· ····················
S 7489 hCoV-19/England/ALDP-B62F7D/2020|EPI_ISL_655360|20 ···················· ···················· ····················· ···················· ···················· ····················
S 7490 hCoV-19/England/ALDP-B5B5B1/2020|EPI_ISL_655361|20 ···················· ···················· ····················· ···················· ···················· ····················
S 7491 hCoV-19/England/ALDP-B62DFB/2020|EPI_ISL_655362|20 ···················· ···················· ····················· ···················· ···················· ····················
S 7492 hCoV-19/England/ALDP-B5B681/2020|EPI_ISL_655363|20 ···················· ···················· ····················· ···················· ···················· ····················
S 7493 hCoV-19/England/ALDP-B62F9B/2020|EPI_ISL_655364|20 ···················· ···················· ····················· ···················· ···················· ····················
S 7494 hCoV-19/England/ALDP-B62C0D/2020|EPI_ISL_655365|20 ···················· ···················· ····················· ···················· ···················· ····················
S 7495 hCoV-19/England/ALDP-B5BB82/2020|EPI_ISL_655367|20 ···················· ·A·················· ····················· ···················· ···················· ···A················
S 7496 hCoV-19/England/ALDP-B5AF3C/2020|EPI_ISL_655368|20 ···················· ···················· ····················· ···················· ···················· ····················
S 7497 hCoV-19/England/ALDP-B5B171/2020|EPI_ISL_655369|20 ···················· ···················· ····················· ···················· ······T············· ····················
S 7498 hCoV-19/England/ALDP-B5B44B/2020|EPI_ISL_655370|20 ···················· ···················· ····················· ···················· ···················· ····················
S 7499 hCoV-19/England/ALDP-B5BD40/2020|EPI_ISL_655371|20 ···················· ···················· ····················· ···················· ···················· ····················
S 7500 hCoV-19/England/ALDP-B5B821/2020|EPI_ISL_655372|20 ···················· ···················· ····················· ···················· ···················· ····················
S 7501 hCoV-19/England/ALDP-B5B593/2020|EPI_ISL_655373|20 ···················· ···················· ····················· ···················· ···················· ····················
S 7502 hCoV-19/England/ALDP-B5B6EB/2020|EPI_ISL_655374|20 ···················· ···················· ····················· ···················· ···················· ····················
S 7503 hCoV-19/England/ALDP-B5B8F4/2020|EPI_ISL_655375|20 ···················· ···················· ····················· ···················· ···················· ····················
S 7504 hCoV-19/England/ALDP-B5BA85/2020|EPI_ISL_655376|20 ···················· ···················· ····················· ···················· ···················· ····················
S 7505 hCoV-19/England/ALDP-B5E67F/2020|EPI_ISL_655377|20 ···················· ···················· ····················· ···················· ···················· ····················
S 7506 hCoV-19/England/ALDP-B5B35D/2020|EPI_ISL_655378|20 ···················· ···················· ····················· ···················· ···················· ····················
S 7507 hCoV-19/England/ALDP-B5B830/2020|EPI_ISL_655380|20 ···················· ···················· ····················· ···················· ···················· ····················
S 7508 hCoV-19/England/ALDP-B5B117/2020|EPI_ISL_655381|20 ···················· ···················· ····················· ···················· ···················· ····················
S 7509 hCoV-19/England/ALDP-B5BBBF/2020|EPI_ISL_655382|20 ···················· ···················· ····················· ···················· ···················· ····················
S 7510 hCoV-19/England/ALDP-B5B4A5/2020|EPI_ISL_655383|20 ···················· ···················· ····················· ···················· ···················· ····················
S 7511 hCoV-19/England/ALDP-B63088/2020|EPI_ISL_655384|20 ···················· ···················· ····················· ···················· ···················· ····················
S 7512 hCoV-19/England/ALDP-B5B029/2020|EPI_ISL_655385|20 ···················· ···················· ····················· ···················· ···················· ····················
S 7513 hCoV-19/England/ALDP-B5BDB9/2020|EPI_ISL_655386|20 ···················· ···················· ····················· ···················· ···················· ····················
S 7514 hCoV-19/England/ALDP-B62FC8/2020|EPI_ISL_655387|20 ···················· ···················· ····················· ···················· ···················· ····················
S 7515 hCoV-19/England/ALDP-B5B2F6/2020|EPI_ISL_655388|20 ···················· ···················· ····················· ···················· ···················· ····················
S 7516 hCoV-19/England/ALDP-B5B8B8/2020|EPI_ISL_655389|20 ···················· ···················· ····················· ···················· ···················· ····················
S 7517 hCoV-19/England/ALDP-B5B205/2020|EPI_ISL_655390|20 ···················· ···················· ····················· ···················· ···················· ····················
S 7518 hCoV-19/England/ALDP-B62CA3/2020|EPI_ISL_655391|20 ···················· ···················· ····················· ···················· ···················· ····················
S 7519 hCoV-19/England/ALDP-B5BA58/2020|EPI_ISL_655392|20 ···················· ···················· ····················· ···················· ···················· ····················
S 7520 hCoV-19/England/ALDP-B63D72/2020|EPI_ISL_655393|20 ···················· ···················· ····················· ···················· ···················· ····················
S 7521 hCoV-19/England/ALDP-B5BA49/2020|EPI_ISL_655394|20 ···················· ···················· ····················· ···················· ···················· ····················
S 7522 hCoV-19/England/ALDP-B5BB46/2020|EPI_ISL_655396|20 ···················· ···················· ····················· ···················· ···················· ····················
S 7523 hCoV-19/England/ALDP-B5B38A/2020|EPI_ISL_655399|20 ···················· ···················· ····················· ···················· ···················· ····················
S 7524 hCoV-19/England/ALDP-B62669/2020|EPI_ISL_656600|20 ···················· ···················· ····················· ···················· ···················· ····················
S 7525 hCoV-19/England/ALDP-B62A8B/2020|EPI_ISL_656601|20 ···················· ···················· ····················· ···················· ···················· ····················
S 7526 hCoV-19/England/ALDP-B62942/2020|EPI_ISL_656602|20 ···················· ···················· ····················· ···················· ···················· ····················
S 7527 hCoV-19/England/ALDP-B638AE/2020|EPI_ISL_656603|20 ···················· ···················· ····················· ···················· ···················· ····················
S 7528 hCoV-19/England/ALDP-B6206B/2020|EPI_ISL_656604|20 ···················· ···················· ····················· ···················· ···················· ····················
S 7529 hCoV-19/England/ALDP-B62380/2020|EPI_ISL_656605|20 ···················· ···················· ····················· ···················· ···················· ····················
S 7530 hCoV-19/England/ALDP-B628EB/2020|EPI_ISL_656606|20 ···················· ···················· ····················· ···················· ···················· ····················
S 7531 hCoV-19/England/ALDP-B639AB/2020|EPI_ISL_656607|20 ···················· ···················· ····················· ···················· ···················· ····················
S 7532 hCoV-19/NorthernIreland/ALDP-B63A11/2020|EPI_ISL_6 ···················· ···················· ····················· ···················· ···················· ····················
S 7533 hCoV-19/England/ALDP-B62960/2020|EPI_ISL_656609|20 ···················· ···················· ····················· ···················· ···················· ····················
S 7534 hCoV-19/England/ALDP-B6353E/2020|EPI_ISL_656610|20 ···················· ···················· ····················· ···················· ···················· ····················

S 7535 hCoV-19/England/ALDP-B6346E/2020|EPI_ISL_656613|20 ···················· ···················· ····················· ···················· ···A················ ····················
S 7537 hCoV-19/England/ALDP-B639D8/2020|EPI_ISL_656615|20 ···················· ···················· ····················· ···················· ···················· ····················
S 7538 hCoV-19/England/ALDP-B6363B/2020|EPI_ISL_656616|20 ···················· ···················· ····················· ···················· ···················· ···A················
S 7539 hCoV-19/England/ALDP-B6396F/2020|EPI_ISL_656617|20 ···················· ···················· ····················· ···················· ···················· ····················
S 7540 hCoV-19/England/ALDP-B63C84/2020|EPI_ISL_656618|20 ···················· ···················· ····················· ···················· ···················· ····················
S 7541 hCoV-19/England/ALDP-B63950/2020|EPI_ISL_656619|20 ···················· ···················· ····················· ···················· ···················· ····················
S 7542 hCoV-19/England/ALDP-B62906/2020|EPI_ISL_656620|20 ···················· ···················· ····················· ···················· ···················· ····················
S 7544 hCoV-19/England/ALDP-B62784/2020|EPI_ISL_656622|20 ···················· ···················· ····················· ···················· ···················· ····················
S 7545 hCoV-19/England/ALDP-B62A30/2020|EPI_ISL_656623|20 ···················· ···················· ····················· ···················· ···················· ····················
S 7546 hCoV-19/England/ALDP-B6257B/2020|EPI_ISL_656624|20 ···················· ···················· ····················· ···················· ···················· ····················
S 7547 hCoV-19/England/ALDP-B63589/2020|EPI_ISL_656625|20 ···················· ···················· ····················· ···················· ···················· ····················
S 7548 hCoV-19/England/ALDP-B63905/2020|EPI_ISL_656628|20 ···················· ···················· ····················· ···················· ···················· ····················
S 7549 hCoV-19/England/ALDP-B63871/2020|EPI_ISL_656629|20 ···················· ···················· ····················· ···················· ···················· ····················
S 7550 hCoV-19/England/ALDP-B62687/2020|EPI_ISL_656630|20 ···················· ···················· ····················· ···················· ······T············· ····················
S 7551 hCoV-19/England/ALDP-B627FD/2020|EPI_ISL_656631|20 ···················· ·A·················· ····················· ···················· ···················· ···A················
S 7553 hCoV-19/England/ALDP-B62432/2020|EPI_ISL_656633|20 ···················· ···················· ····················· ···················· ···················· ····················
S 7554 hCoV-19/England/ALDP-B63431/2020|EPI_ISL_656635|20 ···················· ···················· ····················· ···················· ···A················ ····················
S 7555 hCoV-19/England/ALDP-B62872/2020|EPI_ISL_656647|20 ···················· ···················· ····················· ···················· ···················· ····················
S 7556 hCoV-19/England/ALDP-B6270C/2020|EPI_ISL_656648|20 ···················· ···················· ····················· ···················· ···················· ····················
S 7557 hCoV-19/England/ALDP-B6364A/2020|EPI_ISL_656649|20 ···················· ···················· ····················· ···················· ···················· ····················
S 7558 hCoV-19/England/ALDP-B63792/2020|EPI_ISL_656651|20 ···················· ···················· ····················· ···················· ···················· ····················
S 7559 hCoV-19/England/ALDP-B63307/2020|EPI_ISL_656652|20 ···················· ···················· ····················· ···················· ···················· ····················
S 7560 hCoV-19/England/ALDP-B620E3/2020|EPI_ISL_656653|20 ···················· ···················· ····················· ···················· ···················· ····················
S 7561 hCoV-19/England/ALDP-B626F0/2020|EPI_ISL_656658|20 ···················· ···················· ····················· ···················· ···················· ····················
S 7562 hCoV-19/England/ALDP-B629E8/2020|EPI_ISL_656660|20 ···················· ···················· ····················· ···················· ···················· ····················
S 7563 hCoV-19/England/ALDP-B6271B/2020|EPI_ISL_656661|20 ···················· ···················· ····················· ···················· ···················· ····················
S 7564 hCoV-19/England/ALDP-B6202F/2020|EPI_ISL_656662|20 ···················· ·A·················· ····················· ···················· ···················· ···A················
S 7565 hCoV-19/England/ALDP-B62520/2020|EPI_ISL_656663|20 ···················· ···················· ····················· ···················· ···················· ····················
S 7566 hCoV-19/England/ALDP-B620C5/2020|EPI_ISL_656664|20 ···················· ·A·················· ····················· ···················· ···················· ···A················
S 7567 hCoV-19/England/ALDP-B624F6/2020|EPI_ISL_656665|20 ···················· ···················· ····················· ···················· ···················· ····················
S 7568 hCoV-19/England/ALDP-B6207A/2020|EPI_ISL_656666|20 ···················· ···················· ····················· ···················· ···················· ····················
S 7569 hCoV-19/England/ALDP-B625C6/2020|EPI_ISL_656667|20 ···················· ···················· ····················· ···················· ···················· ····················
S 7570 hCoV-19/England/ALDP-B6338F/2020|EPI_ISL_656668|20 ···················· ···················· ····················· ···················· ···················· ····················
S 7571 hCoV-19/England/ALDP-B63CEE/2020|EPI_ISL_656670|20 ···················· ···················· ····················· ···················· ···················· ····················
S 7572 hCoV-19/England/ALDP-B622DE/2020|EPI_ISL_656671|20 ···················· ···················· ····················· ···················· ···················· ····················
S 7573 hCoV-19/England/ALDP-B62A21/2020|EPI_ISL_656676|20 ···················· ···················· ····················· ···················· ···················· ····················
S 7574 hCoV-19/England/ALDP-B635A7/2020|EPI_ISL_656680|20 ···················· ·A·················· ····················· ···················· ···················· ···A················
S 7575 hCoV-19/England/ALDP-B63237/2020|EPI_ISL_656681|20 ···················· ···················· ····················· ···················· ···················· ····················
S 7576 hCoV-19/England/ALDP-B627DF/2020|EPI_ISL_656682|20 ···················· ···················· ····················· ···················· ···················· ····················
S 7577 hCoV-19/England/ALDP-B632EC/2020|EPI_ISL_656684|20 ···················· ···················· ····················· ···················· ···················· ····················
S 7578 hCoV-19/England/MILK-B56DAF/2020|EPI_ISL_656685|20 ···················· ···················· ····················· ···················· ···················· ····················
S 7579 hCoV-19/England/CAMC-B53FCA/2020|EPI_ISL_656686|20 ···················· ···················· ····················· ···················· ···················· ····················
S 7580 hCoV-19/England/CAMC-B54424/2020|EPI_ISL_656687|20 ···················· ···················· ····················· ···················· ···················· ····················
S 7581 hCoV-19/England/CAMC-B52408/2020|EPI_ISL_656688|20 ···················· ···················· ····················· ···················· ···················· ····················
S 7582 hCoV-19/England/CAMC-B53795/2020|EPI_ISL_656689|20 ···················· ···················· ····················· ···················· ···················· ····················
S 7583 hCoV-19/England/CAMC-B54442/2020|EPI_ISL_656690|20 ···················· ···················· ····················· ···················· ···················· ····················
S 7584 hCoV-19/England/CAMC-B54309/2020|EPI_ISL_656691|20 ···················· ···················· ···········T········· ···················· ···················· ····················
S 7585 hCoV-19/England/CAMC-B54239/2020|EPI_ISL_656692|20 ···················· ·A·················· ····················· ···················· ···················· ···A················
S 7586 hCoV-19/England/CAMC-B526C6/2020|EPI_ISL_656693|20 ···················· ···················· ····················· ···················· ···················· ····················
S 7587 hCoV-19/England/CAMC-B5275A/2020|EPI_ISL_656694|20 ···················· ···················· ····················· ···················· ···················· ····················
S 7588 hCoV-19/England/CAMC-B5267B/2020|EPI_ISL_656699|20 ···················· ···················· ····················· ···················· ···················· ····················
S 7589 hCoV-19/England/MILK-B551A4/2020|EPI_ISL_657901|20 ···················· ···················· ····················· ···················· ···················· ····················
S 7590 hCoV-19/England/CAMC-B520D7/2020|EPI_ISL_657903|20 ···················· ···················· ····················· ···················· ···················· ····················
S 7591 hCoV-19/England/MILK-B54DDE/2020|EPI_ISL_657906|20 ···················· ···················· ····················· ···················· ···················· ····················
S 7592 hCoV-19/England/CAMC-B51C10/2020|EPI_ISL_657907|20 ···················· ···················· ····················· ···················· ···················· ····················
S 7593 hCoV-19/England/CAMC-B511E4/2020|EPI_ISL_657908|20 ···················· ···················· ····················· ···················· ···················· ····················
S 7594 hCoV-19/England/CAMC-B5109C/2020|EPI_ISL_657909|20 ···················· ···················· ····················· ···················· ···················· ····················
S 7595 hCoV-19/England/MILK-B555C6/2020|EPI_ISL_657910|20 ···················· ···················· ····················· ···················· ···················· ····················
S 7596 hCoV-19/England/CAMC-B51FF9/2020|EPI_ISL_657912|20 ···················· ···················· ····················· ···················· ···················· ····················
S 7597 hCoV-19/England/MILK-B54BD4/2020|EPI_ISL_657913|20 ···················· ···················· ····················· ···················· ···················· ····················
S 7598 hCoV-19/England/CAMC-B5124B/2020|EPI_ISL_657914|20 ···················· ···················· ····················· ···················· ···················· ····················
S 7599 hCoV-19/England/MILK-B54BE3/2020|EPI_ISL_657915|20 ···················· ···················· ····················· ···················· ···················· ····················
S 7600 hCoV-19/England/MILK-B553BD/2020|EPI_ISL_657916|20 ···················· ···················· ····················· ···················· ···················· ····················
S 7601 hCoV-19/England/CAMC-B51551/2020|EPI_ISL_657917|20 ···················· ···················· ····················· ···················· ······T············· ····················
S 7602 hCoV-19/England/MILK-B54C77/2020|EPI_ISL_657918|20 ···················· ···················· ····················· ···················· ···················· ····················
S 7603 hCoV-19/England/MILK-B55168/2020|EPI_ISL_657919|20 ···················· ···················· ····················· ···················· ···················· ····················
S 7604 hCoV-19/England/CAMC-B5213E/2020|EPI_ISL_657920|20 ···················· ···················· ····················· ···················· ······T············· ····················
S 7605 hCoV-19/England/MILK-B55292/2020|EPI_ISL_657921|20 ···················· ···················· ····················· ···················· ···················· ····················
S 7606 hCoV-19/England/MILK-B550B6/2020|EPI_ISL_657922|20 ···················· ···················· ····················· ···················· ···················· ····················
S 7607 hCoV-19/England/MILK-B54F23/2020|EPI_ISL_657923|20 ···················· ·A·················· ····················· ···················· ···················· ···A················
S 7608 hCoV-19/England/CAMC-B51E47/2020|EPI_ISL_657924|20 ···················· ···················· ····················· ···················· ···················· ····················
S 7609 hCoV-19/England/MILK-B554C9/2020|EPI_ISL_657925|20 ···················· ·A·················· ····················· ···················· ···················· ···A················
S 7610 hCoV-19/England/CAMC-B51D68/2020|EPI_ISL_657926|20 ···················· ···················· ····················· ···················· ······T············· ····················
S 7611 hCoV-19/England/MILK-B55229/2020|EPI_ISL_657927|20 ···················· ···················· ····················· ···················· ···················· ····················
S 7612 hCoV-19/England/MILK-B54A5F/2020|EPI_ISL_657928|20 ···················· ···················· ····················· ···················· ···················· ····················
S 7613 hCoV-19/England/MILK-B550A7/2020|EPI_ISL_657929|20 ···················· ···················· ····················· ···················· ···················· ····················
S 7614 hCoV-19/England/MILK-B550D4/2020|EPI_ISL_657930|20 ···················· ···················· ····················· ···················· ···················· ····················
S 7615 hCoV-19/England/CAMC-B51F26/2020|EPI_ISL_657931|20 ···················· ···················· ····················· ···················· ···················· ····················
S 7616 hCoV-19/England/MILK-B54CC2/2020|EPI_ISL_657932|20 ···················· ···················· ····················· ···················· ···················· ····················
S 7617 hCoV-19/England/CAMC-B5216B/2020|EPI_ISL_657935|20 ···················· ···················· ····················· ···················· ···················· ····················

| | | | | | | | | |
|---|---|---|---|---|---|---|---|---|
| S | 7618 | hCoV-19/England/CAMC-B510F6/2020\|EPI_ISL_657936\|20 | ···················· | ···················· | ···················· | ···················· | ···················· | ···················· |
| S | 7619 | hCoV-19/England/MILK-B5511D/2020\|EPI_ISL_657938\|20 | ···················· | ···················· | ···················· | ···················· | ···················· | ···················· |
| S | 7620 | hCoV-19/England/CAMC-B51393/2020\|EPI_ISL_657939\|20 | ···················· | ···················· | ···················· | ···················· | ···················· | ···················· |
| S | 7621 | hCoV-19/England/MILK-B55247/2020\|EPI_ISL_657941\|20 | ···················· | ···················· | ···················· | ···················· | ···················· | ···················· |
| S | 7622 | hCoV-19/England/CAMC-B52471/2020\|EPI_ISL_657943\|20 | ···················· | ···················· | ···················· | ···················· | ···················· | ···················· |
| S | 7623 | hCoV-19/England/CAMC-B51E74/2020\|EPI_ISL_657945\|20 | ···················· | ···················· | ···················· | ···················· | ···················· | ···················· |
| S | 7624 | hCoV-19/England/CAMC-B51FBD/2020\|EPI_ISL_657946\|20 | ···················· | ·A·······A·········· | ···················· | ···················· | ···················· | ···················· |
| S | 7625 | hCoV-19/England/MILK-B550E3/2020\|EPI_ISL_657948\|20 | ···················· | ···················· | ···················· | ···················· | ···················· | ···················· |
| S | 7626 | hCoV-19/England/CAMC-B51DB3/2020\|EPI_ISL_657949\|20 | ···················· | ·A·················· | ···················· | ···················· | ···················· | ··A················· |
| S | 7627 | hCoV-19/England/MILK-B54AF5/2020\|EPI_ISL_657950\|20 | ···················· | ···················· | ···················· | ···················· | ···················· | ···················· |
| S | 7628 | hCoV-19/England/CAMC-B512E1/2020\|EPI_ISL_657951\|20 | ···················· | ···················· | ···················· | ···················· | ···················· | ···················· |
| S | 7629 | hCoV-19/England/CAMC-B51C7A/2020\|EPI_ISL_657952\|20 | ···················· | ···················· | ···················· | ···················· | ···················· | ···················· |
| S | 7630 | hCoV-19/England/CAMC-B51FDB/2020\|EPI_ISL_657953\|20 | ···················· | ···················· | ···················· | ···················· | ······T············· | ···················· |
| S | 7631 | hCoV-19/England/MILK-B54AB9/2020\|EPI_ISL_657954\|20 | ···················· | ···················· | ···················· | ···················· | ···················· | ···················· |
| S | 7632 | hCoV-19/England/MILK-B54D1A/2020\|EPI_ISL_657956\|20 | ···················· | ···················· | ···················· | ···················· | ···················· | ···················· |
| S | 7633 | hCoV-19/England/CAMC-B51E56/2020\|EPI_ISL_657957\|20 | ···················· | ···················· | ···················· | ···················· | ···················· | ···················· |
| S | 7634 | hCoV-19/England/MILK-B5555D/2020\|EPI_ISL_657958\|20 | ···················· | ···················· | ···················· | ···················· | ···················· | ···················· |
| S | 7635 | hCoV-19/England/CAMC-B51375/2020\|EPI_ISL_657960\|20 | ···················· | ···················· | ···················· | ···················· | ···················· | ···················· |
| S | 7636 | hCoV-19/England/MILK-B554BA/2020\|EPI_ISL_657961\|20 | ···················· | ·A·················· | ···················· | ···················· | ···················· | ··A················· |
| S | 7637 | hCoV-19/England/MILK-B54C3B/2020\|EPI_ISL_657963\|20 | ···················· | ···················· | ···················· | ···················· | ···················· | ···················· |
| S | 7638 | hCoV-19/England/CAMC-B51CC5/2020\|EPI_ISL_657964\|20 | ···················· | ···················· | ···················· | ···················· | ···················· | ···················· |
| S | 7639 | hCoV-19/England/MILK-B55502/2020\|EPI_ISL_657965\|20 | ···················· | ···················· | ···················· | ···················· | ···················· | ···················· |
| S | 7640 | hCoV-19/England/CAMC-B51D95/2020\|EPI_ISL_657966\|20 | ···················· | ···················· | ···················· | ···················· | ······T············· | ···················· |
| S | 7641 | hCoV-19/England/CAMC-B514BE/2020\|EPI_ISL_657978\|20 | ···················· | ···················· | ···················· | ···················· | ···················· | ···················· |
| S | 7642 | hCoV-19/England/MILK-B54FD8/2020\|EPI_ISL_657982\|20 | ···················· | ···················· | ···················· | ···················· | ···················· | ···················· |
| S | 7643 | hCoV-19/England/CAMC-B51472/2020\|EPI_ISL_657983\|20 | ···················· | ···················· | ···················· | ···················· | ···················· | ···················· |
| S | 7644 | hCoV-19/England/MILK-B54ECC/2020\|EPI_ISL_657984\|20 | ···················· | ···················· | ···················· | ···················· | ···················· | ···················· |
| S | 7645 | hCoV-19/England/CAMC-B5159D/2020\|EPI_ISL_657990\|20 | ···················· | ···················· | ···················· | ···················· | ···················· | ···················· |
| S | 7646 | hCoV-19/England/CAMC-B51F35/2020\|EPI_ISL_657992\|20 | ···················· | ···················· | ···················· | ···················· | ···················· | ···················· |
| S | 7647 | hCoV-19/England/MILK-B55511/2020\|EPI_ISL_657993\|20 | ···················· | ···················· | ···················· | ···················· | ···················· | ···················· |
| S | 7648 | hCoV-19/England/CAMC-B51C01/2020\|EPI_ISL_657994\|20 | ···················· | ·A·················· | ···················· | ···················· | ···················· | ··A················· |
| S | 7649 | hCoV-19/England/MILK-B54AE6/2020\|EPI_ISL_657995\|20 | ···················· | ···················· | ···················· | ···················· | ···················· | ···················· |
| S | 7650 | hCoV-19/England/MILK-B551FF/2020\|EPI_ISL_657996\|20 | ···················· | ···················· | ···················· | ···················· | ···················· | ···················· |
| S | 7651 | hCoV-19/England/CAMC-B51481/2020\|EPI_ISL_657998\|20 | ···················· | ·A·················· | ···················· | ···················· | ···················· | ··A················· |
| S | 7652 | hCoV-19/England/MILK-B55441/2020\|EPI_ISL_657999\|20 | ···················· | ·A·················· | ···················· | ···················· | ···················· | ··A················· |
| S | 7653 | hCoV-19/USA/WI-GMF-32905/2020\|EPI_ISL_661168\|2020- | ···················· | ···················· | ···················· | ···················· | ···················· | ···················· |
| S | 7654 | hCoV-19/USA/IA-GMF-32907/2020\|EPI_ISL_661169\|2020- | ···················· | ···················· | ···················· | ···················· | ···················· | ···················· |
| S | 7655 | hCoV-19/USA/IA-GMF-33129/2020\|EPI_ISL_661170\|2020- | ···················· | ·A·················· | ···················· | ···················· | ···················· | ···················· |
| S | 7656 | hCoV-19/USA/WI-GMF-33162/2020\|EPI_ISL_661171\|2020- | ···················· | ···················· | ···················· | ···················· | ···················· | ···················· |
| S | 7657 | hCoV-19/USA/WI-GMF-33231/2020\|EPI_ISL_661172\|2020- | ···················· | ···················· | ···················· | ···················· | ···················· | ···················· |
| S | 7658 | hCoV-19/USA/WI-GMF-M3001/2020\|EPI_ISL_661173\|2020- | ···················· | ···················· | ···················· | ···················· | ···················· | ···················· |
| S | 7659 | hCoV-19/England/204821174/2020\|EPI_ISL_661175\|2020 | ···················· | ···················· | ···················· | ···················· | ···················· | ···················· |
| S | 7660 | hCoV-19/Serbia/Subotica-198541706/2020\|EPI_ISL_661 | ···················· | ···················· | ···················· | ···················· | ···················· | ···················· |
| S | 7661 | hCoV-19/Serbia/Novi Sad-197031606/2020\|EPI_ISL_661 | ···················· | ···················· | ···················· | ···················· | ···················· | ···················· |
| S | 7662 | hCoV-19/Serbia/Novi Sad-215372606/2020\|EPI_ISL_661 | ···················· | ···················· | ···················· | ···················· | ···················· | ···················· |
| S | 7663 | hCoV-19/Serbia/Backa Palanka-234850407/2020\|EPI_IS | ···················· | ···················· | ···················· | ···················· | ···················· | ···················· |
| S | 7664 | hCoV-19/Serbia/Novi Sad-197021606/2020\|EPI_ISL_661 | ···················· | ···················· | ···················· | ···················· | ···················· | ···················· |
| S | 7665 | hCoV-19/Serbia/Pancevo-210032506/2020\|EPI_ISL_6611 | ···················· | ···················· | ···················· | ···················· | ···················· | ···················· |
| S | 7666 | hCoV-19/Serbia/Novi Sad-198771706/2020\|EPI_ISL_661 | ···················· | ···················· | ···················· | ···················· | ···················· | ···················· |
| S | 7667 | hCoV-19/Serbia/Vrsac-253781007/2020\|EPI_ISL_661186 | ···················· | ···················· | ···················· | ···················· | ···················· | ···················· |
| S | 7668 | hCoV-19/Serbia/Novi Sad-243000607/2020\|EPI_ISL_661 | ···T················ | ···················· | ···················· | ···················· | ···················· | ···················· |
| S | 7669 | hCoV-19/Serbia/Pancevo-215572606/2020\|EPI_ISL_6611 | ···················· | ···················· | ···················· | ···················· | ···················· | ···················· |
| S | 7670 | hCoV-19/Serbia/Kikinda-000201303/2020_\|EPI_ISL_661 | ···················· | ···················· | ···················· | ···················· | ···················· | ···················· |
| S | 7671 | hCoV-19/Serbia/Novi Sad-281762307/2020\|EPI_ISL_661 | ···················· | ·A·················· | ···················· | ···················· | ···················· | ···················· |
| S | 7672 | hCoV-19/Serbia/Novi Sad-209542506/20200\|EPI_ISL_66 | ···················· | ···················· | ···················· | ···················· | ···················· | ···················· |
| S | 7673 | hCoV-19/Serbia/Novi Sad-215042606/2020\|EPI_ISL_661 | ···················· | ···················· | ···················· | ···················· | ···················· | ···················· |
| S | 7674 | hCoV-19/Serbia/Ruma-254521007/2020\|EPI_ISL_661193\| | ···················· | ···················· | ···················· | ···················· | ···················· | ···················· |
| S | 7675 | hCoV-19/Serbia/Novi Sad-283692307/2020\|EPI_ISL_661 | ···················· | ···················· | ···················· | ···················· | ···················· | ···················· |
| S | 7676 | hCoV-19/Serbia/Vrbas-214652606/2020\|EPI_ISL_661195 | ···················· | ···················· | ···················· | ···················· | ···················· | ···················· |
| S | 7677 | hCoV-19/Serbia/Novi Sad-214302606/2020\|EPI_ISL_661 | ···················· | ···················· | ···················· | ···················· | ···················· | ···················· |
| S | 7678 | hCoV-19/Serbia/Vrsac-259991107/2020\|EPI_ISL_661197 | ···················· | ···················· | ···················· | ···················· | ···················· | ···················· |
| S | 7679 | hCoV-19/Serbia/Srbobran-305963007/2020\|EPI_ISL_661 | ···················· | ···················· | ···················· | ···················· | ···················· | ···················· |
| S | 7680 | hCoV-19/Serbia/Vrbas-239470607/2020\|EPI_ISL_661199 | ···················· | ···················· | ···················· | ···················· | ···················· | ···················· |
| S | 7681 | hCoV-19/England/ALDP-B47E9F/2020\|EPI_ISL_649400\|20 | ···················· | ···················· | ···················· | ···················· | ···················· | ···················· |
| S | 7682 | hCoV-19/England/ALDP-B48748/2020\|EPI_ISL_649401\|20 | ···················· | ···················· | ···················· | ···················· | ···················· | ···················· |
| S | 7683 | hCoV-19/England/ALDP-B48775/2020\|EPI_ISL_649402\|20 | ···················· | ···················· | ···················· | ···················· | ···················· | ···················· |
| S | 7684 | hCoV-19/England/ALDP-B47F05/2020\|EPI_ISL_649404\|20 | ···················· | ···················· | ···················· | ···················· | ···················· | ···················· |
| S | 7685 | hCoV-19/England/MILK-B50516/2020\|EPI_ISL_649405\|20 | ···················· | ···················· | ···················· | ···················· | ······T············· | ···················· |
| S | 7686 | hCoV-19/England/ALDP-B48010/2020\|EPI_ISL_649406\|20 | ···················· | ···················· | ···················· | ···················· | ···················· | ···················· |
| S | 7687 | hCoV-19/England/ALDP-B48265/2020\|EPI_ISL_649407\|20 | ···················· | ···················· | ···················· | ···················· | ···················· | ···················· |
| S | 7688 | hCoV-19/England/ALDP-B49334/2020\|EPI_ISL_649408\|20 | ···················· | ···················· | ···················· | ···················· | ···················· | ···················· |
| S | 7689 | hCoV-19/England/ALDP-B48F8C/2020\|EPI_ISL_649409\|20 | ···················· | ···················· | ···················· | ···················· | ···················· | ···················· |
| S | 7690 | hCoV-19/England/ALDP-B49176/2020\|EPI_ISL_649410\|20 | ···················· | ···················· | ···················· | ···················· | ···················· | ···················· |
| S | 7691 | hCoV-19/England/ALDP-B48247/2020\|EPI_ISL_649411\|20 | ···················· | ···················· | ···················· | ···················· | ···················· | ···················· |
| S | 7692 | hCoV-19/England/ALDP-B4848D/2020\|EPI_ISL_649412\|20 | ···················· | ···················· | ···················· | ···················· | ···················· | ···················· |
| S | 7693 | hCoV-19/England/ALDP-B48F9B/2020\|EPI_ISL_649413\|20 | ···················· | ···················· | ···················· | ···················· | ···················· | ···················· |
| S | 7694 | hCoV-19/England/ALDP-B4939E/2020\|EPI_ISL_649414\|20 | ···················· | ···················· | ···················· | ···················· | ···················· | ···················· |
| S | 7695 | hCoV-19/England/ALDP-B4856C/2020\|EPI_ISL_649415\|20 | ···················· | ···················· | ···················· | ···················· | ···················· | ···················· |
| S | 7696 | hCoV-19/England/ALDP-B4807A/2020\|EPI_ISL_649416\|20 | ···················· | ···················· | ···················· | ···················· | ···················· | ···················· |
| S | 7697 | hCoV-19/England/ALDP-B4812C/2020\|EPI_ISL_649417\|20 | ···················· | ···················· | ···················· | ···················· | ···················· | ···················· |

S 7698 hCoV-19/England/ALDP-B483DB/2020|EPI_ISL_649418|20 ···················· ···················· ···················· ···················· ···················· ····················
S 7699 hCoV-19/England/ALDP-B493DA/2020|EPI_ISL_649419|20 ···················· ···················· ···················· ···················· ···················· ····················
S 7700 hCoV-19/England/ALDP-B48511/2020|EPI_ISL_649421|20 ···················· ···················· ···················· ···················· ···················· ····················
S 7701 hCoV-19/England/ALDP-B488BE/2020|EPI_ISL_649422|20 ···················· ···················· ···················· ···················· ···················· ····················
S 7702 hCoV-19/England/ALDP-B486D2/2020|EPI_ISL_649423|20 ···················· ···················· ···················· ···················· ···················· ····················
S 7703 hCoV-19/England/MILK-B50525/2020|EPI_ISL_649424|20 ···················· ···················· ···················· ···················· ···················· ····················
S 7704 hCoV-19/England/ALDP-B4FDE5/2020|EPI_ISL_649426|20 ···················· ···················· ···················· ···················· ···················· ····················
S 7705 hCoV-19/England/ALDP-B49088/2020|EPI_ISL_649427|20 ···················· ·········T·········· ···················· ···················· ···················· ····················
S 7706 hCoV-19/England/ALDP-B491FE/2020|EPI_ISL_649428|20 ···················· ···················· ···················· ···················· ···················· ····················
S 7707 hCoV-19/England/ALDP-B48177/2020|EPI_ISL_649429|20 ···················· ···················· ···················· ···················· ···················· ····················
S 7708 hCoV-19/England/ALDP-B4814A/2020|EPI_ISL_649430|20 ···················· ···················· ···················· ···················· ···················· ····················
S 7709 hCoV-19/England/ALDP-B49370/2020|EPI_ISL_649431|20 ···················· ···················· ···················· ···················· ···················· ····················
S 7710 hCoV-19/England/ALDP-B50D1E/2020|EPI_ISL_649432|20 ···················· ···················· ···················· ···················· ···················· ····················
S 7711 hCoV-19/England/ALDP-B485A8/2020|EPI_ISL_649433|20 ···················· ···················· ···················· ···················· ···················· ····················
S 7712 hCoV-19/England/ALDP-B48E16/2020|EPI_ISL_649435|20 ···················· ···················· ···················· ···················· ···················· ····················
S 7713 hCoV-19/England/ALDP-B4863C/2020|EPI_ISL_649436|20 ···················· ···················· ···················· ···················· ···················· ····················
S 7714 hCoV-19/England/ALDP-B47D29/2020|EPI_ISL_649437|20 ···················· ···················· ···················· ···················· ···················· ····················
S 7715 hCoV-19/England/ALDP-B485C6/2020|EPI_ISL_649438|20 ···················· ···················· ···················· ···················· ···················· ····················
S 7716 hCoV-19/England/ALDP-B47CB3/2020|EPI_ISL_649440|20 ···················· ···················· ···················· ···················· ···················· ····················
S 7717 hCoV-19/England/ALDP-B4803E/2020|EPI_ISL_649441|20 ···················· ···················· ···················· ···················· ···················· ····················
S 7718 hCoV-19/England/ALDP-B48DCE/2020|EPI_ISL_649442|20 ···················· ···················· ···················· ···················· ···················· ····················
S 7719 hCoV-19/England/ALDP-B4864B/2020|EPI_ISL_649443|20 ···················· ···················· ···················· ···················· ···················· ····················
S 7720 hCoV-19/England/ALDP-B47EAE/2020|EPI_ISL_649444|20 ···················· ···················· ···················· ···················· ···················· ····················
S 7721 hCoV-19/England/ALDP-B48186/2020|EPI_ISL_649445|20 ···················· ···················· ···················· ···················· ···················· ····················
S 7722 hCoV-19/England/ALDP-B484BA/2020|EPI_ISL_649446|20 ···················· ···················· ···················· ···················· ···················· ····················
S 7723 hCoV-19/England/ALDP-B49343/2020|EPI_ISL_649447|20 ···················· ···················· ···················· ···················· ···················· ····················
S 7724 hCoV-19/England/ALDP-B47D74/2020|EPI_ISL_649448|20 ···················· ···················· ···················· ···················· ···················· ····················
S 7725 hCoV-19/England/ALDP-B48335/2020|EPI_ISL_649449|20 ···················· ···················· ···················· ···················· ···················· ····················
S 7726 hCoV-19/England/ALDP-B486F0/2020|EPI_ISL_649452|20 ···················· ···················· ···················· ···················· ···················· ····················
S 7727 hCoV-19/England/ALDP-B47F41/2020|EPI_ISL_649453|20 ···················· ···················· ···················· ···················· ···················· ····················
S 7728 hCoV-19/England/ALDP-B47DCF/2020|EPI_ISL_649454|20 ···················· ···················· ···················· ···················· ···················· ····················
S 7729 hCoV-19/England/ALDP-B48678/2020|EPI_ISL_649455|20 ···················· ···················· ···················· ···················· ···················· ····················
S 7730 hCoV-19/England/ALDP-B48669/2020|EPI_ISL_649471|20 ···················· ···················· ···················· ···················· ···················· ····················
S 7731 hCoV-19/England/ALDP-B486A5/2020|EPI_ISL_649472|20 ···················· ···················· ···················· ···················· ···················· ····················
S 7732 hCoV-19/England/ALDP-B48F40/2020|EPI_ISL_649473|20 ···················· ···················· ···················· ···················· ···················· ····················
S 7733 hCoV-19/England/ALDP-B48FF5/2020|EPI_ISL_649474|20 ···················· ···················· ···················· ···················· ···················· ····················
S 7734 hCoV-19/England/ALDP-B48238/2020|EPI_ISL_649475|20 ···················· ···················· ···················· ···················· ···················· ····················
S 7735 hCoV-19/England/ALDP-B4912B/2020|EPI_ISL_649476|20 ···················· ·A·················· ···················· ···················· ···················· ···A················
S 7736 hCoV-19/England/ALDP-B47F23/2020|EPI_ISL_649477|20 ···················· ···················· ···················· ···················· ···················· ····················
S 7737 hCoV-19/England/ALDP-B48E34/2020|EPI_ISL_649478|20 ···················· ···················· ···················· ···················· ···················· ····················
S 7738 hCoV-19/England/MILK-B504EC/2020|EPI_ISL_649479|20 ···················· ···················· ···················· ···················· ···················· ····················
S 7739 hCoV-19/England/ALDP-B4849C/2020|EPI_ISL_649480|20 ···················· ···················· ···················· ···················· ···················· ····················
S 7740 hCoV-19/England/CAMC-B36543/2020|EPI_ISL_649482|20 ···················· ···················· ···················· ···················· ···················· ····················
S 7741 hCoV-19/England/ALDP-B1C543/2020|EPI_ISL_649484|20 ···················· ···················· ···················· ···················· ···················· ····················
S 7742 hCoV-19/England/ALDP-B1F883/2020|EPI_ISL_649485|20 ···················· ···················· ···················· ···················· ···················· ····················
S 7743 hCoV-19/England/ALDP-B24EF2/2020|EPI_ISL_649487|20 ···················· ···················· ···················· ···················· ···················· ····················
S 7744 hCoV-19/England/ALDP-B24645/2020|EPI_ISL_649488|20 ···················· ···················· ···················· ···················· ···················· ····················
S 7745 hCoV-19/England/ALDP-B238AA/2020|EPI_ISL_649489|20 ···················· ···················· ···················· ···················· ···················· ····················
S 7746 hCoV-19/England/ALDP-B1A897/2020|EPI_ISL_649490|20 ···················· ···················· ···················· ···················· ···················· ····················
S 7747 hCoV-19/England/CAMC-B20B86/2020|EPI_ISL_649492|20 ···················· ···················· ···················· ···················· ···················· ····················
S 7748 hCoV-19/England/CAMC-B20843/2020|EPI_ISL_649493|20 ···················· ···················· ···················· ···················· ···················· ····················
S 7749 hCoV-19/Scotland/QEUH-B0F033/2020|EPI_ISL_649494|2 ···················· ···················· ···················· ···················· ···················· ····················
S 7750 hCoV-19/Scotland/QEUH-B12FC7/2020|EPI_ISL_649495|2 ···················· ···················· ···················· ···················· ···················· ····················
S 7751 hCoV-19/Scotland/QEUH-B105C7/2020|EPI_ISL_649497|2 ···················· ···················· ···················· ···················· ···················· ····················
S 7752 hCoV-19/Scotland/QEUH-B0F121/2020|EPI_ISL_649498|2 ···················· ···················· ···················· ···················· ···················· ····················
S 7753 hCoV-19/Scotland/QEUH-B13FA8/2020|EPI_ISL_649499|2 ···················· ···················· ···················· ···················· ···················· ····················
S 7754 hCoV-19/England/CAMB-1B932A/2020|EPI_ISL_651700|20 ···················· ···················· ···················· ···················· ······T············· ····················
S 7755 hCoV-19/England/CAMB-1B9339/2020|EPI_ISL_651701|20 ···················· ···················· ···················· ···················· ···················· ····················
S 7756 hCoV-19/England/CAMB-1B9375/2020|EPI_ISL_651702|20 ···················· ···················· ···················· ···················· ···················· ····················
S 7757 hCoV-19/England/CAMB-1B9384/2020|EPI_ISL_651703|20 ···················· ···················· ···················· ···················· ···················· ····················
S 7758 hCoV-19/England/CAMB-1B9393/2020|EPI_ISL_651704|20 ···················· ···················· ···················· ···················· ···················· ····················
S 7759 hCoV-19/England/CAMB-1B93C0/2020|EPI_ISL_651705|20 ···················· ···················· ···················· ···················· ···················· ····················
S 7760 hCoV-19/England/CAMB-1B93DF/2020|EPI_ISL_651706|20 ···················· ···················· ···················· ···················· ···················· ····················
S 7761 hCoV-19/England/CAMB-1B93EE/2020|EPI_ISL_651707|20 ···················· ···················· ···················· ···················· ···················· ····················
S 7762 hCoV-19/England/CAMB-1B9CF2/2020|EPI_ISL_651708|20 ···················· ···················· ···················· ···················· ···················· ····················
S 7763 hCoV-19/England/CAMB-1B9D4A/2020|EPI_ISL_651709|20 ···················· ···················· ···················· ···················· ···················· ····················
S 7764 hCoV-19/England/CAMB-1B9DA4/2020|EPI_ISL_651710|20 ···················· ···················· ···················· ···················· ···················· ····················
S 7765 hCoV-19/England/CAMB-1B9F80/2020|EPI_ISL_651711|20 ···················· ···················· ···················· ···················· ···················· ····················
S 7766 hCoV-19/England/CAMB-1BA08C/2020|EPI_ISL_651712|20 ···················· ···················· ···················· ···················· ···················· ····················
S 7767 hCoV-19/England/CAMB-1BA0AA/2020|EPI_ISL_651713|20 ···················· ·········T·········· ···················· ···················· ···················· ····················
S 7768 hCoV-19/England/CAMB-1BA12F/2020|EPI_ISL_651714|20 ···················· ···················· ···················· ···················· ···················· ····················
S 7769 hCoV-19/England/CAMC-B23084/2020|EPI_ISL_651715|20 ···················· ···················· ···················· ···················· ···················· ····················
S 7770 hCoV-19/England/CAMC-B230C0/2020|EPI_ISL_651716|20 ···················· ···················· ···················· ···················· ···················· ····················
S 7771 hCoV-19/England/CAMC-B230FD/2020|EPI_ISL_651717|20 ···················· ···················· ···················· ···················· ···················· ····················
S 7772 hCoV-19/England/CAMC-B23109/2020|EPI_ISL_651718|20 ···················· ·A·················· ···················· ···················· ···················· ···A················
S 7773 hCoV-19/England/CAMC-B23136/2020|EPI_ISL_651719|20 ···················· ···················· ···················· ···················· ···················· ····················
S 7774 hCoV-19/England/CAMC-B23145/2020|EPI_ISL_651720|20 ···················· ···················· ···················· ···················· ···················· ····················
S 7775 hCoV-19/England/CAMC-B23172/2020|EPI_ISL_651721|20 ···················· ···················· ···················· ···················· ···················· ····················
S 7776 hCoV-19/England/CAMC-B23181/2020|EPI_ISL_651722|20 ···················· ···················· ···················· ···················· ···················· ····················
S 7777 hCoV-19/England/CAMC-B23215/2020|EPI_ISL_651723|20 ···················· ···················· ···················· ···················· ···················· ····················

| | | | | | | | |
|---|---|---|---|---|---|---|---|
| S | 7778 hCoV-19/England/CAMC-B23224/2020\|EPI_ISL_651724\|20 | ···················· | ···················· | ····················· | ···················· | ···················· | ···················· |
| S | 7779 hCoV-19/England/CAMC-B2327F/2020\|EPI_ISL_651725\|20 | ···················· | ···················· | ····················· | ···················· | ······T············· | ···················· |
| S | 7780 hCoV-19/England/CAMC-B2328E/2020\|EPI_ISL_651726\|20 | ···················· | ···················· | ····················· | ···················· | ···················· | ···················· |
| S | 7781 hCoV-19/England/CAMC-B232BB/2020\|EPI_ISL_651727\|20 | ···················· | ···················· | ····················· | ···················· | ······T············· | ···················· |
| S | 7782 hCoV-19/England/CAMC-B232CA/2020\|EPI_ISL_651728\|20 | ···················· | ···················· | ····················· | ···················· | ······T············· | ···················· |
| S | 7783 hCoV-19/England/CAMC-B234C4/2020\|EPI_ISL_651729\|20 | ···················· | ···················· | ····················· | ···················· | ······T············· | ···················· |
| S | 7784 hCoV-19/England/CAMC-B234E2/2020\|EPI_ISL_651730\|20 | ···················· | ···················· | ····················· | ···················· | ······T············· | ···················· |
| S | 7785 hCoV-19/England/CAMC-B234F1/2020\|EPI_ISL_651731\|20 | ···················· | ···················· | ····················· | ···················· | ···················· | ···················· |
| S | 7786 hCoV-19/England/CAMC-B2350D/2020\|EPI_ISL_651732\|20 | ···················· | ···················· | ····················· | ···················· | ······T············· | ···················· |
| S | 7787 hCoV-19/Scotland/CVR4855/2020\|EPI_ISL_651733\|2020- | ···················· | ···················· | ····················· | ···················· | ···················· | ···················· |
| S | 7788 hCoV-19/Scotland/CVR4864/2020\|EPI_ISL_651734\|2020- | ···················· | ···················· | ····················· | ···················· | ···················· | ···················· |
| S | 7789 hCoV-19/Scotland/CVR4867/2020\|EPI_ISL_651735\|2020- | ···················· | ···················· | ····················· | ···················· | ···················· | ···················· |
| S | 7790 hCoV-19/Scotland/CVR4880/2020\|EPI_ISL_651736\|2020- | ···················· | ···················· | ····················· | ···················· | ···················· | ···················· |
| S | 7791 hCoV-19/Scotland/CVR4886/2020\|EPI_ISL_651737\|2020- | ···················· | ···················· | ····················· | ···················· | ···················· | ···················· |
| S | 7792 hCoV-19/Scotland/CVR4887/2020\|EPI_ISL_651738\|2020- | ···················· | ···················· | ····················· | ···················· | ···················· | ···················· |
| S | 7793 hCoV-19/Scotland/CVR4984/2020\|EPI_ISL_651739\|2020- | ···················· | ···················· | ····················· | ···················· | ···················· | ···················· |
| S | 7794 hCoV-19/Scotland/CVR5288/2020\|EPI_ISL_651740\|2020- | ···················· | ···················· | ····················· | ···················· | ···················· | ···················· |
| S | 7795 hCoV-19/Scotland/CVR5291/2020\|EPI_ISL_651741\|2020- | ···················· | ···················· | ····················· | ···················· | ···················· | ···················· |
| S | 7796 hCoV-19/Scotland/CVR5295/2020\|EPI_ISL_651742\|2020- | ···················· | ···················· | ····················· | ···················· | ···················· | ···················· |
| S | 7797 hCoV-19/Scotland/CVR5299/2020\|EPI_ISL_651743\|2020- | ···················· | ···················· | ····················· | ···················· | ···················· | ···················· |
| S | 7798 hCoV-19/Scotland/CVR5300/2020\|EPI_ISL_651744\|2020- | ···················· | ···················· | ····················· | ···················· | ···················· | ···················· |
| S | 7799 hCoV-19/Scotland/CVR5301/2020\|EPI_ISL_651745\|2020- | ···················· | ···················· | ····················· | ···················· | ···················· | ···················· |
| S | 7800 hCoV-19/Scotland/CVR5303/2020\|EPI_ISL_651746\|2020- | ···················· | ···················· | ····················· | ···················· | ···················· | ···················· |
| S | 7801 hCoV-19/Scotland/CVR5307/2020\|EPI_ISL_651747\|2020- | ···················· | ···················· | ····················· | ···················· | ···················· | ···················· |
| S | 7802 hCoV-19/Scotland/CVR5309/2020\|EPI_ISL_651748\|2020- | ···················· | ···················· | ····················· | ···················· | ···················· | ···················· |
| S | 7803 hCoV-19/Scotland/CVR5310/2020\|EPI_ISL_651749\|2020- | ···················· | ···················· | ····················· | ···················· | ···················· | ···················· |
| S | 7804 hCoV-19/Scotland/CVR5312/2020\|EPI_ISL_651750\|2020- | ···················· | ···················· | ····················· | ···················· | ···················· | ···················· |
| S | 7805 hCoV-19/Scotland/CVR5314/2020\|EPI_ISL_651751\|2020- | ···················· | ···················· | ····················· | ···················· | ···················· | ···················· |
| S | 7806 hCoV-19/Scotland/CVR5315/2020\|EPI_ISL_651752\|2020- | ···················· | ···················· | ····················· | ···················· | ···················· | ···················· |
| S | 7807 hCoV-19/Scotland/CVR5316/2020\|EPI_ISL_651753\|2020- | ···················· | ···················· | ····················· | ···················· | ···················· | ···················· |
| S | 7808 hCoV-19/Scotland/CVR5317/2020\|EPI_ISL_651754\|2020- | ···················· | ···················· | ····················· | ···················· | ···················· | ···················· |
| S | 7809 hCoV-19/Scotland/CVR5321/2020\|EPI_ISL_651755\|2020- | ···················· | ···················· | ····················· | ···················· | ···················· | ···················· |
| S | 7810 hCoV-19/Scotland/CVR5323/2020\|EPI_ISL_651756\|2020- | ···················· | ···················· | ····················· | ···················· | ···················· | ···················· |
| S | 7811 hCoV-19/Scotland/CVR5324/2020\|EPI_ISL_651757\|2020- | ···················· | ···················· | ····················· | ···················· | ···················· | ···················· |
| S | 7812 hCoV-19/Scotland/CVR5325/2020\|EPI_ISL_651758\|2020- | ···················· | ···················· | ····················· | ···················· | ···················· | ···················· |
| S | 7813 hCoV-19/Scotland/CVR5326/2020\|EPI_ISL_651759\|2020- | ···················· | ···················· | ····················· | ···················· | ···················· | ···················· |
| S | 7814 hCoV-19/Scotland/CVR5328/2020\|EPI_ISL_651760\|2020- | ···················· | ···················· | ····················· | ···················· | ···················· | ···················· |
| S | 7815 hCoV-19/Scotland/CVR5329/2020\|EPI_ISL_651761\|2020- | ···················· | ···················· | ····················· | ···················· | ···················· | ···················· |
| S | 7816 hCoV-19/Scotland/CVR5330/2020\|EPI_ISL_651762\|2020- | ···················· | ···················· | ····················· | ···················· | ···················· | ···················· |
| S | 7817 hCoV-19/Scotland/CVR5332/2020\|EPI_ISL_651763\|2020- | ···················· | ···················· | ····················· | ···················· | ···················· | ···················· |
| S | 7818 hCoV-19/Scotland/CVR5333/2020\|EPI_ISL_651764\|2020- | ···················· | ···················· | ····················· | ···················· | ···················· | ···················· |
| S | 7819 hCoV-19/Scotland/CVR5334/2020\|EPI_ISL_651765\|2020- | ···················· | ···················· | ····················· | ···················· | ···················· | ···················· |
| S | 7820 hCoV-19/Scotland/CVR5335/2020\|EPI_ISL_651766\|2020- | ···················· | ···················· | ····················· | ···················· | ···················· | ···················· |
| S | 7821 hCoV-19/Scotland/CVR5346/2020\|EPI_ISL_651769\|2020- | ···················· | ···················· | ····················· | ···················· | ···················· | ···················· |
| S | 7822 hCoV-19/Scotland/CVR5347/2020\|EPI_ISL_651770\|2020- | ···················· | ···················· | ····················· | ···················· | ···················· | ···················· |
| S | 7823 hCoV-19/Scotland/CVR5348/2020\|EPI_ISL_651771\|2020- | ···················· | ···················· | ····················· | ···················· | ···················· | ···················· |
| S | 7824 hCoV-19/Scotland/CVR5351/2020\|EPI_ISL_651772\|2020- | ···················· | ···················· | ····················· | ···················· | ···················· | ···················· |
| S | 7825 hCoV-19/Scotland/CVR5352/2020\|EPI_ISL_651773\|2020- | ···················· | ···················· | ····················· | ···················· | ···················· | ···················· |
| S | 7826 hCoV-19/Scotland/CVR5378/2020\|EPI_ISL_651785\|2020- | ···················· | ···················· | ····················· | ···················· | ···················· | ···················· |
| S | 7827 hCoV-19/Scotland/CVR5379/2020\|EPI_ISL_651786\|2020- | ···················· | ···················· | ····················· | ···················· | ···················· | ···················· |
| S | 7828 hCoV-19/Scotland/CVR5380/2020\|EPI_ISL_651787\|2020- | ···················· | ···················· | ····················· | ···················· | ···················· | ···················· |
| S | 7829 hCoV-19/Scotland/CVR5381/2020\|EPI_ISL_651788\|2020- | ···················· | ···················· | ····················· | ···················· | ···················· | ···················· |
| S | 7830 hCoV-19/Scotland/CVR5382/2020\|EPI_ISL_651789\|2020- | ···················· | ···················· | ····················· | ···················· | ···················· | ···················· |
| S | 7831 hCoV-19/Scotland/CVR5387/2020\|EPI_ISL_651790\|2020- | ···················· | ···················· | ····················· | ···················· | ···················· | ···················· |
| S | 7832 hCoV-19/Scotland/CVR5388/2020\|EPI_ISL_651791\|2020- | ···················· | ···················· | ····················· | ···················· | ···················· | ···················· |
| S | 7833 hCoV-19/Scotland/CVR5389/2020\|EPI_ISL_651792\|2020- | ···················· | ···················· | ····················· | ···················· | ···················· | ···················· |
| S | 7834 hCoV-19/Scotland/CVR5390/2020\|EPI_ISL_651793\|2020- | ···················· | ···················· | ····················· | ···················· | ···················· | ···················· |
| S | 7835 hCoV-19/Scotland/CVR5391/2020\|EPI_ISL_651794\|2020- | ···················· | ···················· | ····················· | ···················· | ···················· | ···················· |
| S | 7836 hCoV-19/Scotland/CVR5392/2020\|EPI_ISL_651795\|2020- | ···················· | ···················· | ····················· | ···················· | ···················· | ···················· |
| S | 7837 hCoV-19/Scotland/CVR5394/2020\|EPI_ISL_651796\|2020- | ···················· | ···················· | ····················· | ···················· | ···················· | ···················· |
| S | 7838 hCoV-19/Scotland/CVR5395/2020\|EPI_ISL_651797\|2020- | ···················· | ···················· | ····················· | ···················· | ···················· | ···················· |
| S | 7839 hCoV-19/Scotland/CVR5398/2020\|EPI_ISL_651798\|2020- | ···················· | ···················· | ····················· | ···················· | ···················· | ···················· |
| S | 7840 hCoV-19/Scotland/CVR5400/2020\|EPI_ISL_651799\|2020- | ···················· | ···················· | ····················· | ···················· | ···················· | ···················· |
| S | 7841 hCoV-19/England/204481953/2020\|EPI_ISL_654007\|2020 | ···················· | ···················· | ····················· | ···················· | ···················· | ···················· |
| S | 7842 hCoV-19/England/204490005/2020\|EPI_ISL_654008\|2020 | ···················· | ···················· | ····················· | ···················· | ···················· | ···················· |
| S | 7843 hCoV-19/England/204490695/2020\|EPI_ISL_654009\|2020 | ···················· | ···················· | ····················· | ···················· | ···················· | ···················· |
| S | 7844 hCoV-19/England/204490706/2020\|EPI_ISL_654010\|2020 | ···················· | ···················· | ····················· | ···················· | ···················· | ···················· |
| S | 7845 hCoV-19/England/204490902/2020\|EPI_ISL_654011\|2020 | ···················· | ···················· | ····················· | ···················· | ···················· | ···················· |
| S | 7846 hCoV-19/England/204490997/2020\|EPI_ISL_654012\|2020 | ···················· | ···················· | ····················· | ···················· | ···················· | ···················· |
| S | 7847 hCoV-19/England/204490998/2020\|EPI_ISL_654013\|2020 | ···················· | ···················· | ····················· | ···················· | ···················· | ···················· |
| S | 7848 hCoV-19/England/204491013/2020\|EPI_ISL_654014\|2020 | ···················· | ···················· | ····················· | ···················· | ···················· | ···················· |
| S | 7849 hCoV-19/England/204491014/2020\|EPI_ISL_654015\|2020 | ···················· | ···················· | ····················· | ···················· | ···················· | ···················· |
| S | 7850 hCoV-19/Tunisia/3913/2020\|EPI_ISL_654016\|2020-03-1 | ···················· | ···················· | ····················· | ···················· | ···················· | ···················· |
| S | 7851 hCoV-19/Tunisia/55262/2020\|EPI_ISL_654017\|2020-11- | ···················· | ···················· | ····················· | ···················· | ···················· | ···················· |
| S | 7852 hCoV-19/England/ALDP-B5B311/2020\|EPI_ISL_655290\|20 | ···················· | ···················· | ····················· | ···················· | ···················· | ···················· |
| S | 7853 hCoV-19/England/ALDP-B630D3/2020\|EPI_ISL_655293\|20 | ···················· | ···················· | ····················· | ···················· | ···················· | ···················· |
| S | 7854 hCoV-19/England/ALDP-B5B663/2020\|EPI_ISL_655295\|20 | ···················· | ···················· | ····················· | ···················· | ······T············· | ···················· |
| S | 7855 hCoV-19/England/ALDP-B5B43C/2020\|EPI_ISL_655296\|20 | ···················· | ···················· | ····················· | ···················· | ···················· | ···················· |
| S | 7856 hCoV-19/England/ALDP-B62E52/2020\|EPI_ISL_655297\|20 | ···················· | ···················· | ····················· | ···················· | ···················· | ···················· |
| S | 7857 hCoV-19/England/ALDP-B5BAC1/2020\|EPI_ISL_655298\|20 | ···················· | ·A·················· | ····················· | ···················· | ···················· | ···A················ |

S 7858 hCoV-19/England/ALDP-B5B618/2020|EPI_ISL_655299|20 ···················· ·A·················· ···················· ···················· ···················· ····················
S 7859 hCoV-19/England/ALDP-B6306A/2020|EPI_ISL_655341|20 ···················· ···················· ···················· ···················· ···················· ····················
S 7860 hCoV-19/England/ALDP-B5B575/2020|EPI_ISL_655342|20 ···················· ···················· ···················· ···················· ···················· ····················
S 7861 hCoV-19/England/ALDP-B5E6AC/2020|EPI_ISL_655343|20 ···················· ···················· ···················· ···················· ···················· ····················
S 7862 hCoV-19/England/ALDP-B5B5FD/2020|EPI_ISL_655347|20 ···················· ···················· ···················· ···················· ···················· ····················
S 7863 hCoV-19/England/ALDP-B62E16/2020|EPI_ISL_655348|20 ···················· ···················· ···················· ···················· ···················· ····················
S 7864 hCoV-19/England/ALDP-B63C1B/2020|EPI_ISL_656669|20 ···················· ···················· ···················· ···················· ···················· ····················
S 7865 hCoV-19/England/ALDP-B63756/2020|EPI_ISL_656672|20 ···················· ···················· ···················· ···················· ···················· ····················
S 7866 hCoV-19/England/ALDP-B63BC3/2020|EPI_ISL_656673|20 ···················· ···················· ···················· ···················· ···················· ····················
S 7867 hCoV-19/England/ALDP-B62353/2020|EPI_ISL_656675|20 ···················· ···················· ···················· ···················· ···················· ····················
S 7868 hCoV-19/England/ALDP-B6246F/2020|EPI_ISL_656678|20 ···················· ···················· ···················· ···················· ···················· ····················
S 7869 hCoV-19/England/ALDP-B62A03/2020|EPI_ISL_656679|20 ···················· ···················· ···················· ···················· ······T············· ····················
S 7870 hCoV-19/England/CAMC-B51F9F/2020|EPI_ISL_657900|20 ···················· ···················· ···················· ···················· ······T············· ····················
S 7871 hCoV-19/USA/WI-GMF-31613/2020|EPI_ISL_661102|2020- ···················· ···················· ···················· ···················· ···················· ····················
S 7872 hCoV-19/USA/WI-GMF-31622/2020|EPI_ISL_661103|2020- ···················· ···················· ···················· ···················· ···················· ····················
S 7873 hCoV-19/USA/WI-GMF-31631/2020|EPI_ISL_661104|2020- ···················· ···················· ···················· ···················· ···················· ····················
S 7874 hCoV-19/USA/WI-GMF-31633/2020|EPI_ISL_661105|2020- ···················· ···················· ···················· ···················· ···················· ····················
S 7875 hCoV-19/USA/WI-GMF-31648/2020|EPI_ISL_661106|2020- ···················· ···················· ···················· ···················· ···················· ····················
S 7876 hCoV-19/USA/IA-GMF-31654/2020|EPI_ISL_661107|2020- ···················· ···················· ···················· ···················· ···················· ····················
S 7877 hCoV-19/USA/WI-GMF-31658/2020|EPI_ISL_661108|2020- ···················· ···················· ···················· ···················· ···················· ····················
S 7878 hCoV-19/USA/WI-GMF-31666/2020|EPI_ISL_661109|2020- ···················· ···················· ···················· ···················· ···················· ····················
S 7879 hCoV-19/USA/WI-GMF-31677/2020|EPI_ISL_661110|2020- ···················· ···················· ···················· ···················· ···················· ····················
S 7880 hCoV-19/USA/WI-GMF-31711/2020|EPI_ISL_661111|2020- ···················· ···················· ···················· ···················· ···················· ····················
S 7881 hCoV-19/USA/WI-GMF-31778/2020|EPI_ISL_661112|2020- ···················· ···················· ···················· ···················· ···················· ····················
S 7882 hCoV-19/USA/WI-GMF-31789/2020|EPI_ISL_661113|2020- ···················· ···················· ···················· ···················· ···················· ····················
S 7883 hCoV-19/USA/WI-GMF-31811/2020|EPI_ISL_661114|2020- ···················· ···················· ···················· ···················· ···················· ····················
S 7884 hCoV-19/USA/WI-GMF-31832/2020|EPI_ISL_661115|2020- ···················· ···················· ···················· ···················· ···················· ····················
S 7885 hCoV-19/USA/WI-GMF-31841/2020|EPI_ISL_661116|2020- ···················· ···················· ···················· ···················· ···················· ····················
S 7886 hCoV-19/USA/WI-GMF-31865/2020|EPI_ISL_661117|2020- ···················· ···················· ···················· ···················· ···················· ····················
S 7887 hCoV-19/USA/WI-GMF-31893/2020|EPI_ISL_661118|2020- ···················· ···················· ···················· ···················· ···················· ····················
S 7888 hCoV-19/USA/WI-GMF-31910/2020|EPI_ISL_661119|2020- ···················· ···················· ···················· ···················· ···················· ····················
S 7889 hCoV-19/USA/WI-GMF-31976/2020|EPI_ISL_661120|2020- ···················· ···················· ···················· ···················· ···················· ····················
S 7890 hCoV-19/USA/WI-GMF-31991/2020|EPI_ISL_661121|2020- ···················· ···················· ···················· ···················· ···················· ····················
S 7891 hCoV-19/USA/WI-GMF-31997/2020|EPI_ISL_661122|2020- ···················· ···················· ···················· ···················· ···················· ····················
S 7892 hCoV-19/USA/IA-GMF-32028/2020|EPI_ISL_661123|2020- ···················· ·A·················· ···················· ···················· ···················· ····················
S 7893 hCoV-19/USA/WI-GMF-32106/2020|EPI_ISL_661124|2020- ···················· ···················· ···················· ···················· ···················· ····················
S 7894 hCoV-19/USA/WI-GMF-32134/2020|EPI_ISL_661125|2020- ···················· ···················· ···················· ···················· ···················· ····················
S 7895 hCoV-19/USA/WI-GMF-32135/2020|EPI_ISL_661126|2020- ···················· ···················· ···················· ···················· ···················· ····················
S 7896 hCoV-19/USA/WI-GMF-32139/2020|EPI_ISL_661127|2020- ···················· ···················· ···················· ···················· ···················· ····················
S 7897 hCoV-19/USA/IA-GMF-32164/2020|EPI_ISL_661128|2020- ···················· ···················· ···················· ···················· ···················· ····················
S 7898 hCoV-19/USA/WI-GMF-32178/2020|EPI_ISL_661129|2020- ···················· ···················· ···················· ···················· ···················· ····················
S 7899 hCoV-19/USA/WI-GMF-32209/2020|EPI_ISL_661130|2020- ···················· ···················· ···················· ···················· ···················· ····················
S 7900 hCoV-19/USA/WI-GMF-32237/2020|EPI_ISL_661131|2020- ···················· ···················· ···················· ···················· ···················· ····················
S 7901 hCoV-19/USA/WI-GMF-32239/2020|EPI_ISL_661132|2020- ···················· ···················· ···················· ···················· ···················· ····················
S 7902 hCoV-19/USA/WI-GMF-32244/2020|EPI_ISL_661133|2020- ···················· ···················· ···················· ···················· ···················· ····················
S 7903 hCoV-19/USA/WI-GMF-32276/2020|EPI_ISL_661134|2020- ···················· ···················· ···················· ···················· ···················· ····················
S 7904 hCoV-19/USA/WI-GMF-32302/2020|EPI_ISL_661135|2020- ···················· ·A·················· ···················· ···················· ···················· ····················
S 7905 hCoV-19/USA/WI-GMF-32349/2020|EPI_ISL_661136|2020- ···················· ···················· ···················· ···················· ···················· ····················
S 7906 hCoV-19/USA/WI-GMF-32351/2020|EPI_ISL_661137|2020- ···················· ···················· ···················· ···················· ···················· ····················
S 7907 hCoV-19/USA/WI-GMF-32355/2020|EPI_ISL_661138|2020- ···················· ···················· ···················· ···················· ···················· ····················
S 7908 hCoV-19/USA/WI-GMF-32358/2020|EPI_ISL_661139|2020- ···················· ···················· ···················· ···················· ···················· ····················
S 7909 hCoV-19/USA/WI-GMF-32385/2020|EPI_ISL_661140|2020- ···················· ···················· ···················· ···················· ···················· ····················
S 7910 hCoV-19/USA/WI-GMF-32389/2020|EPI_ISL_661141|2020- ···················· ···················· ···················· ···················· ···················· ····················
S 7911 hCoV-19/USA/IA-GMF-32395/2020|EPI_ISL_661142|2020- ···················· ···················· ···················· ···················· ···················· ····················
S 7912 hCoV-19/USA/WI-GMF-32399/2020|EPI_ISL_661143|2020- ···················· ···················· ···················· ···················· ···················· ····················
S 7913 hCoV-19/USA/WI-GMF-32403/2020|EPI_ISL_661144|2020- ···················· ···················· ···················· ···················· ···················· ····················
S 7914 hCoV-19/USA/WI-GMF-32409/2020|EPI_ISL_661145|2020- ···················· ·A·················· ···················· ···················· ···················· ····················
S 7915 hCoV-19/USA/WI-GMF-32461/2020|EPI_ISL_661146|2020- ···················· ···················· ···················· ···················· ···················· ····················
S 7916 hCoV-19/USA/WI-GMF-32468/2020|EPI_ISL_661147|2020- ···················· ···················· ···················· ···················· ···················· ····················
S 7917 hCoV-19/USA/WI-GMF-32476/2020|EPI_ISL_661148|2020- ···················· ···················· ···················· ···················· ···················· ····················
S 7918 hCoV-19/USA/WI-GMF-32503/2020|EPI_ISL_661149|2020- ···················· ···················· ···················· ···················· ···················· ····················
S 7919 hCoV-19/USA/WI-GMF-32507/2020|EPI_ISL_661150|2020- ···················· ···················· ···················· ···················· ···················· ····················
S 7920 hCoV-19/USA/WI-GMF-32511/2020|EPI_ISL_661151|2020- ···················· ···················· ···················· ···················· ···················· ····················
S 7921 hCoV-19/USA/WI-GMF-32513/2020|EPI_ISL_661152|2020- ···················· ···················· ···················· ···················· ···················· ····················
S 7922 hCoV-19/USA/WI-GMF-32540/2020|EPI_ISL_661153|2020- ···················· ···················· ···················· ···················· ···················· ····················
S 7923 hCoV-19/USA/WI-GMF-32562/2020|EPI_ISL_661154|2020- ···················· ···················· ···················· ···················· ···················· ····················
S 7924 hCoV-19/USA/WI-GMF-32581/2020|EPI_ISL_661155|2020- ···················· ···················· ···················· ···················· ···················· ····················
S 7925 hCoV-19/USA/IA-GMF-32590/2020|EPI_ISL_661156|2020- ···················· ···················· ···················· ···················· ···················· ····················
S 7926 hCoV-19/USA/WI-GMF-32604/2020|EPI_ISL_661157|2020- ···················· ···················· ···················· ···················· ···················· ····················
S 7927 hCoV-19/USA/WI-GMF-32609/2020|EPI_ISL_661158|2020- ···················· ···················· ···················· ···················· ···················· ····················
S 7928 hCoV-19/USA/WI-GMF-32613/2020|EPI_ISL_661159|2020- ···················· ···················· ···················· ···················· ···················· ····················
S 7929 hCoV-19/USA/MN-GMF-32638/2020|EPI_ISL_661160|2020- ········T··········· ·A·················· ···················· ···················· ···················· ····················
S 7930 hCoV-19/USA/WI-GMF-32640/2020|EPI_ISL_661161|2020- ···················· ···················· ···················· ···················· ···················· ····················
S 7931 hCoV-19/USA/WI-GMF-32678/2020|EPI_ISL_661162|2020- ···················· ···················· ···················· ···················· ···················· ····················
S 7932 hCoV-19/USA/WI-GMF-32685/2020|EPI_ISL_661163|2020- ···················· ···················· ···················· ···················· ···················· ····················
S 7933 hCoV-19/USA/IA-GMF-32700/2020|EPI_ISL_661164|2020- ···················· ·A·················· ···················· ···················· ···················· ····················
S 7934 hCoV-19/USA/WI-GMF-32806/2020|EPI_ISL_661165|2020- ···················· ···················· ···················· ···················· ···················· ····················
S 7935 hCoV-19/USA/WI-GMF-32864/2020|EPI_ISL_661166|2020- ···················· ···················· ···················· ···················· ···················· ····················
S 7936 hCoV-19/USA/MN-GMF-32870/2020|EPI_ISL_661167|2020- ···················· ···················· ···················· ···················· ···················· ····················
S 7937 hCoV-19/England/ALDP-B48F5F/2020|EPI_ISL_649450|20 ···················· ···················· ···················· ···················· ···················· ····················

| | | | | | | | | |
|---|---|---|---|---|---|---|---|---|
| S | 7938 | hCoV-19/England/ALDP-B48757/2020\|EPI_ISL_649456\|20 | ···················· | ···················· | ····················· | ···················· | ···················· | ···················· |
| S | 7939 | hCoV-19/England/ALDP-B50C02/2020\|EPI_ISL_649457\|20 | ···················· | ···················· | ····················· | ···················· | ···················· | ···················· |
| S | 7940 | hCoV-19/England/ALDP-B47FBA/2020\|EPI_ISL_649459\|20 | ···················· | ···················· | ····················· | ···················· | ···················· | ···················· |
| S | 7941 | hCoV-19/England/ALDP-B48423/2020\|EPI_ISL_649460\|20 | ···················· | ···················· | ····················· | ···················· | ···················· | ···················· |
| S | 7942 | hCoV-19/England/ALDP-B4811D/2020\|EPI_ISL_649461\|20 | ···················· | ···················· | ····················· | ···················· | ···················· | ···················· |
| S | 7943 | hCoV-19/England/ALDP-B493CB/2020\|EPI_ISL_649462\|20 | ···················· | ···················· | ····················· | ···················· | ···················· | ···················· |
| S | 7944 | hCoV-19/England/ALDP-B4901F/2020\|EPI_ISL_649463\|20 | ···················· | ···················· | ····················· | ···················· | ···················· | ···················· |
| S | 7945 | hCoV-19/England/ALDP-B4853F/2020\|EPI_ISL_649464\|20 | ···················· | ···················· | ····················· | ···················· | ···················· | ···················· |
| S | 7946 | hCoV-19/England/ALDP-B4938F/2020\|EPI_ISL_649465\|20 | ···················· | ···················· | ····················· | ···················· | ···················· | ···················· |
| S | 7947 | hCoV-19/England/ALDP-B48E25/2020\|EPI_ISL_649466\|20 | ···················· | ···················· | ····················· | ···················· | ···················· | ···················· |
| S | 7948 | hCoV-19/England/ALDP-B47F8D/2020\|EPI_ISL_649467\|20 | ···················· | ···················· | ····················· | ···················· | ···················· | ···················· |
| S | 7949 | hCoV-19/England/ALDP-B487A2/2020\|EPI_ISL_649468\|20 | ···················· | ·A·················· | ····················· | ···················· | ···················· | ··A················· |
| S | 7950 | hCoV-19/England/ALDP-B4872A/2020\|EPI_ISL_649469\|20 | ···················· | ···················· | ····················· | ···················· | ···················· | ···················· |
| S | 7951 | hCoV-19/Scotland/CVR5338/2020\|EPI_ISL_651767\|2020- | ···················· | ···················· | ····················· | ···················· | ···················· | ···················· |
| S | 7952 | hCoV-19/Scotland/CVR5342/2020\|EPI_ISL_651768\|2020- | ···················· | ···················· | ····················· | ···················· | ···················· | ···················· |
| S | 7953 | hCoV-19/Scotland/CVR5353/2020\|EPI_ISL_651774\|2020- | ···················· | ···················· | ····················· | ···················· | ···················· | ···················· |
| S | 7954 | hCoV-19/Scotland/CVR5355/2020\|EPI_ISL_651775\|2020- | ···················· | ···················· | ····················· | ···················· | ···················· | ···················· |
| S | 7955 | hCoV-19/Scotland/CVR5356/2020\|EPI_ISL_651776\|2020- | ···················· | ···················· | ····················· | ···················· | ···················· | ···················· |
| S | 7956 | hCoV-19/Scotland/CVR5358/2020\|EPI_ISL_651777\|2020- | ···················· | ···················· | ····················· | ···················· | ···················· | ···················· |
| S | 7957 | hCoV-19/Scotland/CVR5359/2020\|EPI_ISL_651778\|2020- | ···················· | ···················· | ····················· | ···················· | ···················· | ···················· |
| S | 7958 | hCoV-19/Scotland/CVR5360/2020\|EPI_ISL_651779\|2020- | ···················· | ···················· | ····················· | ···················· | ···················· | ···················· |
| S | 7959 | hCoV-19/Scotland/CVR5366/2020\|EPI_ISL_651780\|2020- | ···················· | ···················· | ····················· | ···················· | ···················· | ···················· |
| S | 7960 | hCoV-19/Scotland/CVR5367/2020\|EPI_ISL_651781\|2020- | ···················· | ···················· | ····················· | ···················· | ···················· | ···················· |
| S | 7961 | hCoV-19/Scotland/CVR5370/2020\|EPI_ISL_651782\|2020- | ···················· | ···················· | ····················· | ···················· | ···················· | ···················· |
| S | 7962 | hCoV-19/Scotland/CVR5374/2020\|EPI_ISL_651783\|2020- | ···················· | ···················· | ····················· | ···················· | ···················· | ···················· |
| S | 7963 | hCoV-19/Scotland/CVR5377/2020\|EPI_ISL_651784\|2020- | ···················· | ···················· | ····················· | ···················· | ···················· | ···················· |
| S | 7964 | hCoV-19/England/QEUH-B6B9FE/2020\|EPI_ISL_655200\|20 | ···················· | ···················· | ····················· | ···················· | ····T··············· | ···················· |
| S | 7965 | hCoV-19/England/QEUH-B6C957/2020\|EPI_ISL_655201\|20 | ···················· | ···················· | ····················· | ···················· | ···················· | ···················· |
| S | 7966 | hCoV-19/England/QEUH-B66823/2020\|EPI_ISL_655202\|20 | ···················· | ···················· | ····················· | ···················· | ······T············· | ···················· |
| S | 7967 | hCoV-19/England/QEUH-B66AA5/2020\|EPI_ISL_655203\|20 | ···················· | ···················· | ····················· | ···················· | ···················· | ···················· |
| S | 7968 | hCoV-19/England/QEUH-B68593/2020\|EPI_ISL_655204\|20 | ···················· | ···················· | ····················· | ···················· | ···················· | ···················· |
| S | 7969 | hCoV-19/England/QEUH-B6C896/2020\|EPI_ISL_655205\|20 | ···················· | ···················· | ····················· | ···················· | ···················· | ···················· |
| S | 7970 | hCoV-19/England/QEUH-B6ACC9/2020\|EPI_ISL_655228\|20 | ···················· | ···················· | ····················· | ···················· | ···················· | ···················· |
| S | 7971 | hCoV-19/England/QEUH-B69C8E/2020\|EPI_ISL_655230\|20 | ···················· | ···················· | ····················· | ···················· | ···················· | ···················· |
| S | 7972 | hCoV-19/England/QEUH-B687BB/2020\|EPI_ISL_655231\|20 | ···················· | ···················· | ····················· | ···················· | ···················· | ···················· |
| S | 7973 | hCoV-19/England/QEUH-B67D8D/2020\|EPI_ISL_655232\|20 | ···················· | ···················· | ····················· | ···················· | ···················· | ···················· |
| S | 7974 | hCoV-19/England/QEUH-B68C9E/2020\|EPI_ISL_655233\|20 | ···················· | ···················· | ····················· | ···················· | ···················· | ···················· |
| S | 7975 | hCoV-19/England/QEUH-B6B9EF/2020\|EPI_ISL_655234\|20 | ···················· | ···················· | ····················· | ···················· | ···················· | ···················· |
| S | 7976 | hCoV-19/England/QEUH-B6C975/2020\|EPI_ISL_655235\|20 | ···················· | ···················· | ····················· | ···················· | ···················· | ···················· |
| S | 7977 | hCoV-19/England/QEUH-B69C51/2020\|EPI_ISL_655236\|20 | ···················· | ···················· | ····················· | ···················· | ···················· | ···················· |
| S | 7978 | hCoV-19/England/QEUH-B68A58/2020\|EPI_ISL_655237\|20 | ···················· | ···················· | ····················· | ···················· | ···················· | ···················· |
| S | 7979 | hCoV-19/England/QEUH-B6BE95/2020\|EPI_ISL_655238\|20 | ···················· | ···················· | ····················· | ···················· | ···················· | ···················· |
| S | 7980 | hCoV-19/England/QEUH-B68A67/2020\|EPI_ISL_655240\|20 | ···················· | ···················· | ····················· | ···················· | ···················· | ···················· |
| S | 7981 | hCoV-19/England/QEUH-B6C474/2020\|EPI_ISL_655241\|20 | ···················· | ···················· | ····················· | ···················· | ···················· | ···················· |
| S | 7982 | hCoV-19/England/QEUH-B695FC/2020\|EPI_ISL_655245\|20 | ···················· | ···················· | ····················· | ···················· | ···················· | ···················· |
| S | 7983 | hCoV-19/England/QEUH-B6BA19/2020\|EPI_ISL_655250\|20 | ···················· | ···················· | ····················· | ···················· | ···················· | ···················· |
| S | 7984 | hCoV-19/England/QEUH-B67CEA/2020\|EPI_ISL_655251\|20 | ···················· | ···················· | ····················· | ···················· | ···················· | ···················· |
| S | 7985 | hCoV-19/England/QEUH-B66780/2020\|EPI_ISL_655252\|20 | ···················· | ···················· | ····················· | ···················· | ···················· | ···················· |
| S | 7986 | hCoV-19/England/QEUH-B68A49/2020\|EPI_ISL_655253\|20 | ···················· | ···················· | ····················· | ···················· | ···················· | ···················· |
| S | 7987 | hCoV-19/England/QEUH-B6A2A9/2020\|EPI_ISL_655254\|20 | ···················· | ···················· | ····················· | ···················· | ···················· | ···················· |
| S | 7988 | hCoV-19/England/QEUH-B69529/2020\|EPI_ISL_655255\|20 | ···················· | ···················· | ····················· | ···················· | ···················· | ···················· |
| S | 7989 | hCoV-19/England/QEUH-B66A78/2020\|EPI_ISL_655256\|20 | ···················· | ···················· | ····················· | ···················· | ···················· | ···················· |
| S | 7990 | hCoV-19/England/QEUH-B6C948/2020\|EPI_ISL_655257\|20 | ···················· | ···················· | ····················· | ···················· | ···················· | ···················· |
| S | 7991 | hCoV-19/England/QEUH-B69C42/2020\|EPI_ISL_655258\|20 | ···················· | ···················· | ····················· | ···················· | ···················· | ···················· |
| S | 7992 | hCoV-19/England/QEUH-B6C8F0/2020\|EPI_ISL_655259\|20 | ···················· | ···················· | ····················· | ···················· | ······T············· | ···················· |
| S | 7993 | hCoV-19/England/QEUH-B6827E/2020\|EPI_ISL_655262\|20 | ···················· | ···················· | ····················· | ···················· | ···················· | ···················· |
| S | 7994 | hCoV-19/England/QEUH-B6888B/2020\|EPI_ISL_655264\|20 | ···················· | ···················· | ····················· | ···················· | ···················· | ···················· |
| S | 7995 | hCoV-19/England/QEUH-B6C043/2020\|EPI_ISL_655265\|20 | ···················· | ···················· | ····················· | ···················· | ···················· | ···········A········ |
| S | 7996 | hCoV-19/England/QEUH-B66A1E/2020\|EPI_ISL_655266\|20 | ···················· | ···················· | ····················· | ···················· | ···················· | ···················· |
| S | 7997 | hCoV-19/England/QEUH-B69E88/2020\|EPI_ISL_655267\|20 | ···················· | ···················· | ····················· | ···················· | ······T············· | ···················· |
| S | 7998 | hCoV-19/England/QEUH-B69E97/2020\|EPI_ISL_655268\|20 | ···················· | ···················· | ····················· | ···················· | ···················· | ···················· |
| S | 7999 | hCoV-19/England/QEUH-B67D32/2020\|EPI_ISL_655269\|20 | ···················· | ···················· | ····················· | ···················· | ···················· | ···················· |
| S | 8000 | hCoV-19/England/QEUH-B687D9/2020\|EPI_ISL_655270\|20 | ···················· | ···················· | ····················· | ···················· | ···················· | ···················· |
| S | 8001 | hCoV-19/England/QEUH-B6C483/2020\|EPI_ISL_655272\|20 | ···················· | ···················· | ····················· | ···················· | ···················· | ···················· |
| S | 8002 | hCoV-19/England/ALDP-B5BAA3/2020\|EPI_ISL_655273\|20 | ···················· | ···················· | ····················· | ···················· | ···················· | ···················· |
| S | 8003 | hCoV-19/England/ALDP-B62F5F/2020\|EPI_ISL_655274\|20 | ···················· | ···················· | ····················· | ···················· | ···A················ | ···················· |
| S | 8004 | hCoV-19/England/ALDP-B62F6E/2020\|EPI_ISL_655275\|20 | ···················· | ···················· | ····················· | ···················· | ···················· | ···················· |
| S | 8005 | hCoV-19/England/ALDP-B62D37/2020\|EPI_ISL_655276\|20 | ···················· | ···················· | ····················· | ···················· | ···················· | ···················· |
| S | 8006 | hCoV-19/England/ALDP-B5B232/2020\|EPI_ISL_655277\|20 | ···················· | ·A·················· | ····················· | ···················· | ···················· | ··A················· |
| S | 8007 | hCoV-19/England/ALDP-B5B162/2020\|EPI_ISL_655278\|20 | ···················· | ···················· | ····················· | ···················· | ···················· | ···················· |
| S | 8008 | hCoV-19/England/ALDP-B62EE9/2020\|EPI_ISL_655279\|20 | ···················· | ···················· | ····················· | ···················· | ···················· | ···················· |
| S | 8009 | hCoV-19/England/ALDP-B5B95B/2020\|EPI_ISL_655280\|20 | ···················· | ···················· | ····················· | ···················· | ···················· | ···················· |
| S | 8010 | hCoV-19/England/ALDP-B5B7BB/2020\|EPI_ISL_655281\|20 | ···················· | ···················· | ····················· | ···················· | ···················· | ···················· |
| S | 8011 | hCoV-19/England/ALDP-B5BBEC/2020\|EPI_ISL_655282\|20 | ···················· | ···················· | ····················· | ···················· | ···················· | ···················· |
| S | 8012 | hCoV-19/England/ALDP-B5E6D9/2020\|EPI_ISL_655283\|20 | ···················· | ·A·················· | ····················· | ···················· | ···················· | ···················· |
| S | 8013 | hCoV-19/England/ALDP-B63CDF/2020\|EPI_ISL_655284\|20 | ···················· | ···················· | ····················· | ···················· | ···················· | ···················· |
| S | 8014 | hCoV-19/England/ALDP-B5BC34/2020\|EPI_ISL_655286\|20 | ···················· | ···················· | ····················· | ···················· | ···················· | ···················· |
| S | 8015 | hCoV-19/England/ALDP-B5B34E/2020\|EPI_ISL_655287\|20 | ···················· | ···················· | ····················· | ···················· | ···················· | ···················· |
| S | 8016 | hCoV-19/England/ALDP-B5B3B7/2020\|EPI_ISL_655288\|20 | ···················· | ···················· | ····················· | ···················· | ···················· | ···················· |
| S | 8017 | hCoV-19/England/ALDP-B62E8F/2020\|EPI_ISL_655289\|20 | ···················· | ···················· | ····················· | ···················· | ···················· | ···················· |

| | | | | | | | | |
|---|---|---|---|---|---|---|---|---|
| S | 8018 | hCoV-19/England/ALDP-B638EA/2020\|EPI_ISL_656500\|20 | ···················· | ·A·················· | ···················· | ···················· | ···················· | ··A················· |
| S | 8019 | hCoV-19/England/ALDP-B63A8A/2020\|EPI_ISL_656501\|20 | ···················· | ···················· | ···················· | ···················· | ···················· | ···················· |
| S | 8020 | hCoV-19/England/ALDP-B627B1/2020\|EPI_ISL_656502\|20 | ···················· | ···················· | ···················· | ···················· | ···················· | ···················· |
| S | 8021 | hCoV-19/England/ALDP-B63264/2020\|EPI_ISL_656503\|20 | ···················· | ···················· | ···················· | ···················· | ···················· | ···················· |
| S | 8022 | hCoV-19/England/ALDP-B632CE/2020\|EPI_ISL_656505\|20 | ···················· | ···················· | ···················· | ···················· | ···················· | ···················· |
| S | 8023 | hCoV-19/England/ALDP-B6354D/2020\|EPI_ISL_656506\|20 | ···················· | ···················· | ···················· | ···················· | ···················· | ···················· |
| S | 8024 | hCoV-19/England/ALDP-B638F9/2020\|EPI_ISL_656507\|20 | ···················· | ···················· | ···················· | ···················· | ···················· | ···················· |
| S | 8025 | hCoV-19/England/ALDP-B633CB/2020\|EPI_ISL_656508\|20 | ···················· | ···················· | ···················· | ···················· | ···················· | ···················· |
| S | 8026 | hCoV-19/England/ALDP-B629D9/2020\|EPI_ISL_656510\|20 | ···················· | ···················· | ···················· | ···················· | ···················· | ···················· |
| S | 8027 | hCoV-19/England/ALDP-B62089/2020\|EPI_ISL_656511\|20 | ···················· | ···················· | ···················· | ···················· | ·····T·············· | ···················· |
| S | 8028 | hCoV-19/England/ALDP-B61FBA/2020\|EPI_ISL_656512\|20 | ···················· | ···················· | ···················· | ···················· | ···················· | ···················· |
| S | 8029 | hCoV-19/England/ALDP-B63AC6/2020\|EPI_ISL_656514\|20 | ···················· | ···················· | ···················· | ···················· | ···················· | ···················· |
| S | 8030 | hCoV-19/England/ALDP-B6347D/2020\|EPI_ISL_656515\|20 | ···················· | ···················· | ···················· | ···················· | ··A················· | ···················· |
| S | 8031 | hCoV-19/England/ALDP-B62863/2020\|EPI_ISL_656516\|20 | ···················· | ···················· | ···················· | ···················· | ···················· | ···················· |
| S | 8032 | hCoV-19/England/ALDP-B63BD2/2020\|EPI_ISL_656517\|20 | ···················· | ···················· | ···················· | ···················· | ···················· | ···················· |
| S | 8033 | hCoV-19/England/ALDP-B638BD/2020\|EPI_ISL_656518\|20 | ···················· | ···················· | ···················· | ···················· | ···················· | ···················· |
| S | 8034 | hCoV-19/England/ALDP-B61F41/2020\|EPI_ISL_656519\|20 | ···················· | ···················· | ···················· | ···················· | ···················· | ···················· |
| S | 8036 | hCoV-19/England/ALDP-B63AD5/2020\|EPI_ISL_656521\|20 | ···················· | ···················· | ···················· | ···················· | ···················· | ···················· |
| S | 8037 | hCoV-19/England/ALDP-B634AA/2020\|EPI_ISL_656522\|20 | ···················· | ···················· | ···················· | ···················· | ···················· | ···················· |
| S | 8038 | hCoV-19/England/ALDP-B6389F/2020\|EPI_ISL_656523\|20 | ···················· | ···················· | ···················· | ···················· | ···················· | ···················· |
| S | 8039 | hCoV-19/England/ALDP-B63413/2020\|EPI_ISL_656524\|20 | ···················· | ···················· | ···················· | ···················· | ···················· | ···················· |
| S | 8040 | hCoV-19/England/ALDP-B632BF/2020\|EPI_ISL_656525\|20 | ···················· | ···················· | ···················· | ···················· | ···················· | ···················· |
| S | 8041 | hCoV-19/England/ALDP-B62344/2020\|EPI_ISL_656526\|20 | ···················· | ···················· | ···················· | ···················· | ···················· | ···················· |
| S | 8042 | hCoV-19/England/ALDP-B626D2/2020\|EPI_ISL_656527\|20 | ···················· | ···················· | ···················· | ···················· | ···················· | ···················· |
| S | 8043 | hCoV-19/England/ALDP-B63BA5/2020\|EPI_ISL_656528\|20 | ···················· | ···················· | ···················· | ···················· | ···················· | ···················· |
| S | 8044 | hCoV-19/England/ALDP-B63A5D/2020\|EPI_ISL_656529\|20 | ···················· | ···················· | ···················· | ···················· | ···················· | ···················· |
| S | 8045 | hCoV-19/England/ALDP-B63B0F/2020\|EPI_ISL_656530\|20 | ···················· | ···················· | ···················· | ···················· | ···················· | ···················· |
| S | 8046 | hCoV-19/England/ALDP-B6260F/2020\|EPI_ISL_656531\|20 | ···················· | ···················· | ···················· | ···················· | ···················· | ···················· |
| S | 8047 | hCoV-19/England/ALDP-B63B3C/2020\|EPI_ISL_656532\|20 | ···················· | ···················· | ···················· | ···················· | ···················· | ···················· |
| S | 8048 | hCoV-19/England/ALDP-B6239F/2020\|EPI_ISL_656533\|20 | ···················· | ···················· | ···················· | ···················· | ···················· | ···················· |
| S | 8050 | hCoV-19/England/ALDP-B633AD/2020\|EPI_ISL_656559\|20 | ···················· | ·A·················· | ···················· | ···················· | ···················· | ··A················· |
| S | 8051 | hCoV-19/England/ALDP-B62AE5/2020\|EPI_ISL_656560\|20 | ···················· | ···················· | ···················· | ···················· | ···················· | ···················· |
| S | 8052 | hCoV-19/England/ALDP-B6356B/2020\|EPI_ISL_656561\|20 | ···················· | ···················· | ···················· | ···················· | ···················· | ···················· |
| S | 8053 | hCoV-19/England/ALDP-B633E9/2020\|EPI_ISL_656562\|20 | ···················· | ···················· | ···················· | ···················· | ···················· | ···················· |
| S | 8054 | hCoV-19/England/ALDP-B6399C/2020\|EPI_ISL_656563\|20 | ···················· | ···················· | ···················· | ···················· | ···················· | ···················· |
| S | 8055 | hCoV-19/England/ALDP-B63817/2020\|EPI_ISL_656564\|20 | ···················· | ···················· | ···················· | ···················· | ···················· | ···················· |
| S | 8056 | hCoV-19/England/ALDP-B62890/2020\|EPI_ISL_656565\|20 | ···················· | ···················· | ···················· | ···················· | ···················· | ···················· |
| S | 8057 | hCoV-19/England/ALDP-B627EE/2020\|EPI_ISL_656566\|20 | ···················· | ·A·················· | ···················· | ···················· | ···················· | ··A················· |
| S | 8058 | hCoV-19/England/ALDP-B62177/2020\|EPI_ISL_656567\|20 | ···················· | ···················· | ···················· | ···················· | ···················· | ···················· |
| S | 8059 | hCoV-19/England/ALDP-B61FE7/2020\|EPI_ISL_656568\|20 | ···················· | ···················· | ···················· | ···················· | ···················· | ···················· |
| S | 8060 | hCoV-19/England/ALDP-B62371/2020\|EPI_ISL_656569\|20 | ···················· | ···················· | ···················· | ···················· | ···················· | ···················· |
| S | 8061 | hCoV-19/England/ALDP-B6398D/2020\|EPI_ISL_656570\|20 | ···················· | ···················· | ···················· | ···················· | ···················· | ···················· |
| S | 8062 | hCoV-19/England/ALDP-B6203E/2020\|EPI_ISL_656571\|20 | ···················· | ···················· | ···················· | ···················· | ···················· | ···················· |
| S | 8063 | hCoV-19/England/ALDP-B62757/2020\|EPI_ISL_656572\|20 | ···················· | ···················· | ···················· | ···················· | ···················· | ···················· |
| S | 8064 | hCoV-19/England/ALDP-B6362C/2020\|EPI_ISL_656581\|20 | ···················· | ·A·················· | ···················· | ···················· | ···················· | ··A················· |
| S | 8065 | hCoV-19/England/ALDP-B6320A/2020\|EPI_ISL_656583\|20 | ···················· | ···················· | ···················· | ···················· | ···················· | ···················· |
| S | 8066 | hCoV-19/England/ALDP-B633F8/2020\|EPI_ISL_656584\|20 | ···················· | ···················· | ···················· | ···················· | ···················· | ···················· |
| S | 8067 | hCoV-19/England/ALDP-B63914/2020\|EPI_ISL_656585\|20 | ···················· | ···················· | ···················· | ···················· | ·····T·············· | ···················· |
| S | 8068 | hCoV-19/England/ALDP-B63CB1/2020\|EPI_ISL_656586\|20 | ···················· | ···················· | ···················· | ···················· | ···················· | ···················· |
| S | 8069 | hCoV-19/England/ALDP-B635C5/2020\|EPI_ISL_656587\|20 | ···················· | ···················· | ···················· | ···················· | ···················· | ···················· |
| S | 8071 | hCoV-19/England/ALDP-B62924/2020\|EPI_ISL_656590\|20 | ···················· | ···················· | ···················· | ···················· | ···················· | ···················· |
| S | 8072 | hCoV-19/England/ALDP-B628DC/2020\|EPI_ISL_656591\|20 | ···········T········ | ···················· | ···················· | ···················· | ···················· | ···················· |
| S | 8073 | hCoV-19/England/ALDP-B635B6/2020\|EPI_ISL_656592\|20 | ···················· | ···················· | ···················· | ···················· | ···················· | ···················· |
| S | 8074 | hCoV-19/England/ALDP-B62775/2020\|EPI_ISL_656593\|20 | ···················· | ···················· | ···················· | ···················· | ···················· | ···················· |
| S | 8075 | hCoV-19/England/ALDP-B63AB7/2020\|EPI_ISL_656595\|20 | ···················· | ···················· | ···················· | ···················· | ···················· | ···················· |
| S | 8076 | hCoV-19/England/ALDP-B61F7E/2020\|EPI_ISL_656596\|20 | ···················· | ···················· | ···················· | ···················· | ···················· | ···················· |
| S | 8077 | hCoV-19/England/ALDP-B6352F/2020\|EPI_ISL_656597\|20 | ···················· | ···················· | ···················· | ···················· | ···················· | ···················· |
| S | 8078 | hCoV-19/England/ALDP-B628FA/2020\|EPI_ISL_656599\|20 | ···················· | ···················· | ···················· | ···················· | ···················· | ···················· |
| S | 8079 | hCoV-19/England/CAMC-B51515/2020\|EPI_ISL_657801\|20 | ···················· | ···················· | ···················· | ···················· | ···················· | ···················· |
| S | 8080 | hCoV-19/England/CAMC-B5130C/2020\|EPI_ISL_657802\|20 | ···················· | ·A·······A·········· | ···················· | ···················· | ···················· | ···················· |
| S | 8081 | hCoV-19/England/CAMC-B511B7/2020\|EPI_ISL_657803\|20 | ···················· | ···················· | ···················· | ···················· | ···················· | ···················· |
| S | 8082 | hCoV-19/England/CAMC-B51D4A/2020\|EPI_ISL_657804\|20 | ···················· | ···················· | ···················· | ···················· | ···················· | ···················· |
| S | 8083 | hCoV-19/England/CAMC-B52110/2020\|EPI_ISL_657805\|20 | ···················· | ·A·················· | ···················· | ···················· | ···················· | ··A················· |
| S | 8084 | hCoV-19/England/CAMC-B5121E/2020\|EPI_ISL_657806\|20 | ···················· | ···················· | ···················· | ···················· | ·····T·············· | ···················· |
| S | 8085 | hCoV-19/England/CAMC-B51269/2020\|EPI_ISL_657807\|20 | ···················· | ···················· | ···················· | ···················· | ···················· | ···················· |
| S | 8086 | hCoV-19/England/MILK-B5546F/2020\|EPI_ISL_657808\|20 | ···················· | ···················· | ···················· | ···················· | ···················· | ···················· |
| S | 8087 | hCoV-19/England/CAMC-B51F62/2020\|EPI_ISL_657809\|20 | ···················· | ···················· | ···················· | ···················· | ···················· | ···················· |
| S | 8088 | hCoV-19/England/CAMC-B514AF/2020\|EPI_ISL_657810\|20 | ···················· | ···················· | ···················· | ···················· | ···················· | ···················· |
| S | 8089 | hCoV-19/England/CAMC-B51EA1/2020\|EPI_ISL_657813\|20 | ···················· | ···················· | ···················· | ···················· | ···················· | ···················· |
| S | 8090 | hCoV-19/England/MILK-B54B6B/2020\|EPI_ISL_657814\|20 | ···················· | ···················· | ···················· | ···················· | ···················· | ···················· |
| S | 8091 | hCoV-19/England/CAMC-B51454/2020\|EPI_ISL_657815\|20 | ···················· | ···················· | ···················· | ···················· | ···················· | ···················· |
| S | 8092 | hCoV-19/England/CAMC-B51DE0/2020\|EPI_ISL_657816\|20 | ···················· | ···················· | ···················· | ···················· | ···················· | ···················· |
| S | 8093 | hCoV-19/England/CAMC-B51E83/2020\|EPI_ISL_657817\|20 | ···················· | ···················· | ···················· | ···················· | ···················· | ···················· |
| S | 8094 | hCoV-19/England/CAMC-B51533/2020\|EPI_ISL_657818\|20 | ···················· | ···················· | ···················· | ···················· | ···················· | ···················· |
| S | 8095 | hCoV-19/England/MILK-B55283/2020\|EPI_ISL_657819\|20 | ···················· | ···················· | ···················· | ···················· | ···················· | ···················· |
| S | 8096 | hCoV-19/England/MILK-B54E35/2020\|EPI_ISL_657822\|20 | ···················· | ···················· | ···················· | ···················· | ···················· | ···················· |
| S | 8097 | hCoV-19/England/MILK-B54D74/2020\|EPI_ISL_657823\|20 | ···················· | ···················· | ···················· | ···················· | ···················· | ···················· |
| S | 8098 | hCoV-19/England/CAMC-B51348/2020\|EPI_ISL_657824\|20 | ···················· | ···················· | ···················· | ···················· | ···················· | ···················· |
| S | 8099 | hCoV-19/England/CAMC-B5108D/2020\|EPI_ISL_657825\|20 | ···················· | ···················· | ···················· | ···················· | ···················· | ···················· |
| S | 8100 | hCoV-19/England/CAMC-B51EDE/2020\|EPI_ISL_657826\|20 | ···················· | ···················· | ···················· | ···················· | ···················· | ···················· |

| | | | | | | | | |
|---|---|---|---|---|---|---|---|---|
| S | 8101 | hCoV-19/England/MILK-B54D56/2020\|EPI_ISL_657827\|20 | ···················· | ···················· | ···················· | ···················· | ···················· | ···················· |
| S | 8102 | hCoV-19/England/MILK-B55432/2020\|EPI_ISL_657829\|20 | ···················· | ···················· | ···················· | ···················· | ···················· | ···················· |
| S | 8103 | hCoV-19/England/MILK-B54F14/2020\|EPI_ISL_657830\|20 | ···················· | ···················· | ···················· | ···················· | ···················· | ···················· |
| S | 8104 | hCoV-19/England/CAMC-B5131B/2020\|EPI_ISL_657831\|20 | ···················· | ···················· | ···················· | ···················· | ···················· | ···················· |
| S | 8105 | hCoV-19/England/MILK-B55238/2020\|EPI_ISL_657832\|20 | ···················· | ···················· | ···················· | ···················· | ···················· | ···················· |
| S | 8106 | hCoV-19/England/CAMC-B51C4D/2020\|EPI_ISL_657833\|20 | ···················· | ···················· | ···················· | ···················· | ···················· | ···················· |
| S | 8107 | hCoV-19/England/CAMC-B51199/2020\|EPI_ISL_657834\|20 | ···················· | ···················· | ···················· | ···················· | ···················· | ···················· |
| S | 8108 | hCoV-19/England/MILK-B55405/2020\|EPI_ISL_657835\|20 | ···················· | ···················· | ···················· | ···················· | ···················· | ···················· |
| S | 8109 | hCoV-19/England/MILK-B54A6E/2020\|EPI_ISL_657836\|20 | ···················· | ···················· | ···················· | ···················· | ···················· | ···················· |
| S | 8110 | hCoV-19/England/CAMC-B51357/2020\|EPI_ISL_657837\|20 | ···················· | ···················· | ···················· | ···················· | ···················· | ···················· |
| S | 8111 | hCoV-19/England/CAMC-B51E38/2020\|EPI_ISL_657839\|20 | ···················· | ···················· | ···················· | ···················· | ···················· | ···················· |
| S | 8112 | hCoV-19/England/CAMC-B51D0E/2020\|EPI_ISL_657841\|20 | ···················· | ···················· | ···················· | ···················· | ···················· | ···················· |
| S | 8113 | hCoV-19/England/MILK-B54E17/2020\|EPI_ISL_657843\|20 | ···················· | ···················· | ···················· | ···················· | ···················· | ···················· |
| S | 8114 | hCoV-19/England/CAMC-B512D2/2020\|EPI_ISL_657845\|20 | ···················· | ···················· | ···················· | ···················· | ···················· | ···················· |
| S | 8115 | hCoV-19/England/CAMC-B51384/2020\|EPI_ISL_657846\|20 | ···················· | ···················· | ···················· | ···················· | ···················· | ···················· |
| S | 8116 | hCoV-19/England/MILK-B54FAB/2020\|EPI_ISL_657850\|20 | ···················· | ···················· | ···················· | ···················· | ···················· | ···················· |
| S | 8117 | hCoV-19/England/CAMC-B51EFC/2020\|EPI_ISL_657852\|20 | ···················· | ···················· | ···················· | ···················· | ···················· | ···················· |
| S | 8118 | hCoV-19/England/CAMC-B5123C/2020\|EPI_ISL_657853\|20 | ···················· | ···················· | ···················· | ···················· | ···················· | ···················· |
| S | 8119 | hCoV-19/England/MILK-B54F41/2020\|EPI_ISL_657854\|20 | ···················· | ···················· | ···················· | ···················· | ·····T·············· | ···················· |
| S | 8120 | hCoV-19/England/CAMC-B51D3B/2020\|EPI_ISL_657855\|20 | ···················· | ···················· | ···················· | ···················· | ·····T·············· | ···················· |
| S | 8121 | hCoV-19/England/CAMC-B52514/2020\|EPI_ISL_657857\|20 | ···················· | ···················· | ···················· | ···················· | ···················· | ···················· |
| S | 8122 | hCoV-19/England/MILK-B550F2/2020\|EPI_ISL_657858\|20 | ···················· | ···················· | ···················· | ···················· | ···················· | ···················· |
| S | 8123 | hCoV-19/England/MILK-B54D65/2020\|EPI_ISL_657859\|20 | ···················· | ···················· | ···················· | ···················· | ···················· | ···················· |
| S | 8124 | hCoV-19/England/CAMC-B51F53/2020\|EPI_ISL_657860\|20 | ···················· | ···················· | ···················· | ···················· | ···················· | ···················· |
| S | 8125 | hCoV-19/England/MILK-B54F7E/2020\|EPI_ISL_657862\|20 | ···················· | ···················· | ···················· | ···················· | ···················· | ···················· |
| S | 8126 | hCoV-19/England/CAMC-B510E7/2020\|EPI_ISL_657863\|20 | ···················· | ···················· | ···················· | ···················· | ···················· | ···················· |
| S | 8127 | hCoV-19/England/MILK-B54EBD/2020\|EPI_ISL_657865\|20 | ···················· | ···················· | ···················· | ···················· | ···················· | ···················· |
| S | 8128 | hCoV-19/England/CAMC-B51463/2020\|EPI_ISL_657866\|20 | ···················· | ···················· | ···················· | ···················· | ···················· | ···················· |
| S | 8129 | hCoV-19/England/CAMC-B52198/2020\|EPI_ISL_657867\|20 | ···················· | ···················· | ···················· | ···················· | ···················· | ···················· |
| S | 8130 | hCoV-19/England/CAMC-B51278/2020\|EPI_ISL_657890\|20 | ···················· | ···················· | ···················· | ···················· | ···················· | ···················· |
| S | 8131 | hCoV-19/England/MILK-B54BB6/2020\|EPI_ISL_657892\|20 | ···················· | ···················· | ···················· | ···················· | ···················· | ···················· |
| S | 8132 | hCoV-19/England/CAMC-B520C8/2020\|EPI_ISL_657893\|20 | ···················· | ···················· | ···················· | ···················· | ···················· | ···················· |
| S | 8133 | hCoV-19/England/MILK-B55335/2020\|EPI_ISL_657894\|20 | ···················· | ···················· | ···················· | ···················· | ···················· | ···················· |
| S | 8134 | hCoV-19/England/CAMC-B515D9/2020\|EPI_ISL_657895\|20 | ···················· | ···················· | ···················· | ···················· | ···················· | ···················· |
| S | 8135 | hCoV-19/England/CAMC-B52031/2020\|EPI_ISL_657896\|20 | ···················· | ···················· | ···················· | ···················· | ···················· | ···················· |
| S | 8136 | hCoV-19/England/CAMC-B513A2/2020\|EPI_ISL_657897\|20 | ···················· | ···················· | ···················· | ···················· | ···················· | ···················· |
| S | 8137 | hCoV-19/England/MILK-B55326/2020\|EPI_ISL_657898\|20 | ···················· | ···················· | ···················· | ···················· | ···················· | ···················· |
| S | 8138 | hCoV-19/USA/WI-GMF-30719/2020\|EPI_ISL_661047\|2020- | ···················· | ·A·················· | ···················· | ···················· | ···················· | ···················· |
| S | 8139 | hCoV-19/USA/MN-GMF-30721/2020\|EPI_ISL_661048\|2020- | ···················· | ···················· | ···················· | ···················· | ···················· | ···················· |
| S | 8140 | hCoV-19/USA/WI-GMF-30775/2020\|EPI_ISL_661049\|2020- | ···················· | ···················· | ···················· | ···················· | ···················· | ···················· |
| S | 8141 | hCoV-19/USA/MN-GMF-30776/2020\|EPI_ISL_661050\|2020- | ···················· | ···················· | ···················· | ···················· | ···················· | ···················· |
| S | 8142 | hCoV-19/USA/WI-GMF-30778/2020\|EPI_ISL_661051\|2020- | ···················· | ···················· | ···················· | ···················· | ···················· | ···················· |
| S | 8143 | hCoV-19/USA/MN-GMF-30812/2020\|EPI_ISL_661052\|2020- | ···················· | ···················· | ···················· | ···················· | ···················· | ···················· |
| S | 8144 | hCoV-19/USA/WI-GMF-30814/2020\|EPI_ISL_661053\|2020- | ···················· | ···················· | ···················· | ···················· | ···················· | ···················· |
| S | 8145 | hCoV-19/USA/WI-GMF-30815/2020\|EPI_ISL_661054\|2020- | ···················· | ···················· | ···················· | ···················· | ···················· | ···················· |
| S | 8146 | hCoV-19/USA/MN-GMF-30816/2020\|EPI_ISL_661055\|2020- | ···················· | ···················· | ···················· | ···················· | ···················· | ···················· |
| S | 8147 | hCoV-19/USA/WI-GMF-30820/2020\|EPI_ISL_661056\|2020- | ···················· | ···················· | ···················· | ···················· | ···················· | ···················· |
| S | 8148 | hCoV-19/USA/WI-GMF-30823/2020\|EPI_ISL_661057\|2020- | ···················· | ···················· | ···················· | ···················· | ···················· | ···················· |
| S | 8149 | hCoV-19/USA/WI-GMF-30826/2020\|EPI_ISL_661058\|2020- | ···················· | ··C················· | ···················· | ···················· | ···················· | ···················· |
| S | 8150 | hCoV-19/USA/WI-GMF-30863/2020\|EPI_ISL_661059\|2020- | ···················· | ···················· | ···················· | ···················· | ···················· | ···················· |
| S | 8151 | hCoV-19/USA/MN-GMF-30871/2020\|EPI_ISL_661060\|2020- | ···················· | ···················· | ···················· | ···················· | ···················· | ···················· |
| S | 8152 | hCoV-19/USA/WI-GMF-30874/2020\|EPI_ISL_661061\|2020- | ···················· | ···················· | ···················· | ···················· | ···················· | ···················· |
| S | 8153 | hCoV-19/USA/WI-GMF-30885/2020\|EPI_ISL_661062\|2020- | ···················· | ···················· | ···················· | ···················· | ···················· | ···················· |
| S | 8154 | hCoV-19/USA/WI-GMF-30889/2020\|EPI_ISL_661063\|2020- | ···················· | ···················· | ···················· | ···················· | ···················· | ···················· |
| S | 8155 | hCoV-19/USA/WI-GMF-30893/2020\|EPI_ISL_661064\|2020- | ···················· | ···················· | ···················· | ···················· | ···················· | ···················· |
| S | 8156 | hCoV-19/USA/WI-GMF-30903/2020\|EPI_ISL_661065\|2020- | ···················· | ···················· | ···················· | ···················· | ···················· | ···················· |
| S | 8157 | hCoV-19/USA/WI-GMF-30919/2020\|EPI_ISL_661066\|2020- | ···················· | ···················· | ···················· | ···················· | ···················· | ···················· |
| S | 8158 | hCoV-19/USA/WI-GMF-30924/2020\|EPI_ISL_661067\|2020- | ···················· | ···················· | ···················· | ···················· | ···················· | ···················· |
| S | 8159 | hCoV-19/USA/WI-GMF-30939/2020\|EPI_ISL_661068\|2020- | ···················· | ···················· | ···················· | ···················· | ···················· | ···················· |
| S | 8160 | hCoV-19/USA/WI-GMF-30992/2020\|EPI_ISL_661069\|2020- | ···················· | ···················· | ···················· | ···················· | ···················· | ···················· |
| S | 8161 | hCoV-19/USA/WI-GMF-31071/2020\|EPI_ISL_661070\|2020- | ···················· | ···················· | ···················· | ···················· | ···················· | ···················· |
| S | 8162 | hCoV-19/USA/WI-GMF-31078/2020\|EPI_ISL_661071\|2020- | ···················· | ···················· | ···················· | ···················· | ···················· | ···················· |
| S | 8163 | hCoV-19/USA/WI-GMF-31086/2020\|EPI_ISL_661072\|2020- | ···················· | ···················· | ···················· | ···················· | ···················· | ···················· |
| S | 8164 | hCoV-19/USA/WI-GMF-31111/2020\|EPI_ISL_661073\|2020- | ···················· | ···················· | ···················· | ···················· | ···················· | ···················· |
| S | 8165 | hCoV-19/USA/WI-GMF-31134/2020\|EPI_ISL_661074\|2020- | ···················· | ···················· | ···················· | ···················· | ···················· | ···················· |
| S | 8166 | hCoV-19/USA/MN-GMF-31147/2020\|EPI_ISL_661075\|2020- | ···················· | ···················· | ···················· | ···················· | ···················· | ···················· |
| S | 8167 | hCoV-19/USA/MN-GMF-31153/2020\|EPI_ISL_661076\|2020- | ···················· | ···················· | ···················· | ···················· | ···················· | ···················· |
| S | 8168 | hCoV-19/USA/WI-GMF-31213/2020\|EPI_ISL_661077\|2020- | ···················· | ···················· | ···················· | ···················· | ···················· | ···················· |
| S | 8169 | hCoV-19/USA/WI-GMF-31217/2020\|EPI_ISL_661078\|2020- | ·······T············ | ···················· | ···················· | ···················· | ···················· | ···················· |
| S | 8170 | hCoV-19/USA/MN-GMF-31256/2020\|EPI_ISL_661079\|2020- | ···················· | ···················· | ···················· | ···················· | ···················· | ···················· |
| S | 8171 | hCoV-19/USA/WI-GMF-31258/2020\|EPI_ISL_661080\|2020- | ···················· | ···················· | ···················· | ···················· | ···················· | ···················· |
| S | 8172 | hCoV-19/USA/WI-GMF-31262/2020\|EPI_ISL_661081\|2020- | ···················· | ···················· | ···················· | ···················· | ···················· | ···················· |
| S | 8173 | hCoV-19/USA/IA-GMF-31330/2020\|EPI_ISL_661082\|2020- | ···················· | ···················· | ···················· | ···················· | ···················· | ···················· |
| S | 8174 | hCoV-19/USA/WI-GMF-31338/2020\|EPI_ISL_661083\|2020- | ···················· | ···················· | ···················· | ···················· | ···················· | ···················· |
| S | 8175 | hCoV-19/USA/WI-GMF-31357/2020\|EPI_ISL_661084\|2020- | ···················· | ·A·················· | ···················· | ···················· | ···················· | ···················· |
| S | 8176 | hCoV-19/USA/MN-GMF-31369/2020\|EPI_ISL_661085\|2020- | ···················· | ···················· | ···················· | ···················· | ···················· | ···················· |
| S | 8177 | hCoV-19/USA/WI-GMF-31372/2020\|EPI_ISL_661086\|2020- | ···················· | ···················· | ···················· | ···················· | ···················· | ···················· |
| S | 8178 | hCoV-19/USA/MN-GMF-31401/2020\|EPI_ISL_661087\|2020- | ···················· | ···················· | ···················· | ···················· | ···················· | ···················· |
| S | 8179 | hCoV-19/USA/WI-GMF-31410/2020\|EPI_ISL_661088\|2020- | ···················· | ···················· | ···················· | ···················· | ···················· | ···················· |
| S | 8180 | hCoV-19/USA/IA-GMF-31421/2020\|EPI_ISL_661089\|2020- | ···················· | ···················· | ···················· | ···················· | ···················· | ···················· |

| | | | | | | | | |
|---|---|---|---|---|---|---|---|---|
| S | 8181 | hCoV-19/USA/WI-GMF-31430/2020\|EPI_ISL_661090\|2020- | ···················· | ···················· | ···················· | ···················· | ···················· | ···················· |
| S | 8182 | hCoV-19/USA/WI-GMF-31443/2020\|EPI_ISL_661091\|2020- | ···················· | ···················· | ···················· | ···················· | ···················· | ···················· |
| S | 8183 | hCoV-19/USA/MN-GMF-31445/2020\|EPI_ISL_661092\|2020- | ···················· | ·A·················· | ···················· | ···················· | ···················· | ···················· |
| S | 8184 | hCoV-19/USA/MN-GMF-31482/2020\|EPI_ISL_661093\|2020- | ···················· | ···················· | ···················· | ···················· | ···················· | ···················· |
| S | 8185 | hCoV-19/USA/WI-GMF-31490/2020\|EPI_ISL_661094\|2020- | ···················· | ···················· | ···················· | ···················· | ···················· | ···················· |
| S | 8186 | hCoV-19/USA/WI-GMF-31492/2020\|EPI_ISL_661095\|2020- | ···················· | ···················· | ···················· | ···················· | ···················· | ···················· |
| S | 8187 | hCoV-19/USA/MN-GMF-31559/2020\|EPI_ISL_661096\|2020- | ···················· | ···················· | ···················· | ···················· | ···················· | ···················· |
| S | 8188 | hCoV-19/USA/WI-GMF-31576/2020\|EPI_ISL_661097\|2020- | ···················· | ···················· | ···················· | ···················· | ···················· | ···················· |
| S | 8189 | hCoV-19/USA/WI-GMF-31577/2020\|EPI_ISL_661098\|2020- | ···················· | ···················· | ···················· | ···················· | ···················· | ···················· |
| S | 8190 | hCoV-19/USA/WI-GMF-31578/2020\|EPI_ISL_661099\|2020- | ···················· | ···················· | ···················· | ···················· | ···················· | ···················· |
| S | 8191 | hCoV-19/USA/WI-GMF-31600/2020\|EPI_ISL_661100\|2020- | ···················· | ···················· | ···················· | ···················· | ···················· | ···················· |
| S | 8192 | hCoV-19/USA/WI-GMF-31611/2020\|EPI_ISL_661101\|2020- | ···················· | ···················· | ···················· | ···················· | ···················· | ···················· |
| S | 8193 | hCoV-19/England/ALDP-B47E53/2020\|EPI_ISL_649300\|20 | ···················· | ···················· | ···················· | ···················· | ···················· | ···················· |
| S | 8194 | hCoV-19/England/ALDP-B4802F/2020\|EPI_ISL_649301\|20 | ···················· | ···················· | ···················· | ···················· | ···················· | ···················· |
| S | 8195 | hCoV-19/England/ALDP-B47F7E/2020\|EPI_ISL_649302\|20 | ···················· | ···················· | ···················· | ···················· | ···················· | ···················· |
| S | 8196 | hCoV-19/England/ALDP-B483AE/2020\|EPI_ISL_649304\|20 | ···················· | ···················· | ···················· | ···················· | ···················· | ···················· |
| S | 8197 | hCoV-19/England/ALDP-B4FF76/2020\|EPI_ISL_649305\|20 | ···················· | ···················· | ···················· | ···················· | ···················· | ···················· |
| S | 8198 | hCoV-19/England/ALDP-B482FC/2020\|EPI_ISL_649306\|20 | ···················· | ···················· | ···················· | ···················· | ···················· | ···················· |
| S | 8199 | hCoV-19/England/ALDP-B48098/2020\|EPI_ISL_649307\|20 | ···················· | ···················· | ···················· | ···················· | ···················· | ···················· |
| S | 8200 | hCoV-19/England/ALDP-B47FE7/2020\|EPI_ISL_649308\|20 | ···················· | ···················· | ···················· | ···················· | ···················· | ···················· |
| S | 8201 | hCoV-19/England/ALDP-B4F9F0/2020\|EPI_ISL_649309\|20 | ···················· | ···················· | ···················· | ···················· | ···················· | ···················· |
| S | 8202 | hCoV-19/England/ALDP-B48520/2020\|EPI_ISL_649311\|20 | ···················· | ···················· | ···················· | ···················· | ···················· | ···················· |
| S | 8203 | hCoV-19/England/ALDP-AC0F8D/2020\|EPI_ISL_649312\|20 | ···················· | ···················· | ···················· | ···················· | ···················· | ···················· |
| S | 8204 | hCoV-19/England/ALDP-B48809/2020\|EPI_ISL_649313\|20 | ···················· | ···················· | ···················· | ···················· | ···················· | ···················· |
| S | 8205 | hCoV-19/England/ALDP-B49361/2020\|EPI_ISL_649315\|20 | ···················· | ···················· | ···················· | ···················· | ···················· | ···················· |
| S | 8206 | hCoV-19/England/ALDP-B481A4/2020\|EPI_ISL_649316\|20 | ···················· | ···················· | ···················· | ···················· | ···················· | ···················· |
| S | 8207 | hCoV-19/England/ALDP-B47D56/2020\|EPI_ISL_649317\|20 | ···················· | ···················· | ···················· | ···················· | ···················· | ···················· |
| S | 8208 | hCoV-19/England/ALDP-B480E3/2020\|EPI_ISL_649318\|20 | ···················· | ···················· | ···················· | ···················· | ···················· | ···A················ |
| S | 8209 | hCoV-19/England/ALDP-B49431/2020\|EPI_ISL_649319\|20 | ···················· | ·········T·········· | ···················· | ···················· | ···················· | ···················· |
| S | 8210 | hCoV-19/England/MILK-B50570/2020\|EPI_ISL_649320\|20 | ···················· | ···················· | ···················· | ···················· | ···················· | ···················· |
| S | 8211 | hCoV-19/England/ALDP-B507A7/2020\|EPI_ISL_649321\|20 | ···················· | ···················· | ···················· | ···················· | ···················· | ···················· |
| S | 8212 | hCoV-19/England/ALDP-B490F1/2020\|EPI_ISL_649322\|20 | ···················· | ···················· | ···················· | ···················· | ···················· | ···················· |
| S | 8213 | hCoV-19/England/ALDP-B48739/2020\|EPI_ISL_649323\|20 | ···················· | ···················· | ···················· | ···················· | ···················· | ···················· |
| S | 8214 | hCoV-19/England/ALDP-B47FAB/2020\|EPI_ISL_649324\|20 | ···················· | ···················· | ···················· | ···················· | ···················· | ···················· |
| S | 8215 | hCoV-19/England/ALDP-B47E26/2020\|EPI_ISL_649325\|20 | ···················· | ···················· | ···················· | ···················· | ···················· | ···················· |
| S | 8216 | hCoV-19/England/ALDP-B4913A/2020\|EPI_ISL_649326\|20 | ···················· | ···················· | ···················· | ···················· | ···················· | ···················· |
| S | 8217 | hCoV-19/England/ALDP-B487DF/2020\|EPI_ISL_649327\|20 | ···················· | ···················· | ···················· | ···················· | ···················· | ···················· |
| S | 8218 | hCoV-19/England/ALDP-B492A0/2020\|EPI_ISL_649328\|20 | ···················· | ···················· | ···················· | ···················· | ···················· | ···················· |
| S | 8219 | hCoV-19/England/ALDP-B490E2/2020\|EPI_ISL_649329\|20 | ···················· | ···················· | ···················· | ···················· | ···················· | ···················· |
| S | 8220 | hCoV-19/England/ALDP-B48793/2020\|EPI_ISL_649390\|20 | ···················· | ···················· | ···················· | ···················· | ···················· | ···················· |
| S | 8221 | hCoV-19/England/ALDP-B48317/2020\|EPI_ISL_649391\|20 | ···················· | ···················· | ···················· | ···················· | ···················· | ···················· |
| S | 8222 | hCoV-19/England/ALDP-B47F9C/2020\|EPI_ISL_649392\|20 | ···················· | ···················· | ···················· | ···················· | ···················· | ···················· |
| S | 8223 | hCoV-19/England/ALDP-B480C5/2020\|EPI_ISL_649393\|20 | ···················· | ···················· | ···················· | ···················· | ···················· | ···················· |
| S | 8224 | hCoV-19/England/ALDP-B48195/2020\|EPI_ISL_649394\|20 | ···················· | ···················· | ···················· | ···················· | ···················· | ···················· |
| S | 8225 | hCoV-19/England/ALDP-B490D3/2020\|EPI_ISL_649395\|20 | ···················· | ·········T·········· | ···················· | ···················· | ···················· | ···················· |
| S | 8226 | hCoV-19/England/ALDP-B490C4/2020\|EPI_ISL_649396\|20 | ···················· | ···················· | ···················· | ···················· | ···················· | ···················· |
| S | 8227 | hCoV-19/England/ALDP-B485B7/2020\|EPI_ISL_649397\|20 | ···················· | ···················· | ···················· | ···················· | ···················· | ···················· |
| S | 8228 | hCoV-19/England/ALDP-B48766/2020\|EPI_ISL_649398\|20 | ···················· | ···················· | ···················· | ···················· | ···················· | ···················· |
| S | 8229 | hCoV-19/England/ALDP-B47E08/2020\|EPI_ISL_649399\|20 | ···················· | ···················· | ···················· | ···················· | ···················· | ···················· |
| S | 8230 | hCoV-19/England/BIRM-66EC2/2020\|EPI_ISL_651600\|202 | ···················· | ···················· | ···················· | ···················· | ···················· | ···················· |
| S | 8231 | hCoV-19/England/BIRM-66ED1/2020\|EPI_ISL_651601\|202 | ···················· | ·A·················· | ···················· | ···················· | ···················· | ···A················ |
| S | 8235 | hCoV-19/England/BIRM-67034/2020\|EPI_ISL_651605\|202 | ···················· | ···················· | ···················· | ···················· | ···················· | ···················· |
| S | 8236 | hCoV-19/England/BIRM-67043/2020\|EPI_ISL_651606\|202 | ···················· | ···················· | ···················· | ···················· | ···················· | ···················· |
| S | 8238 | hCoV-19/England/BIRM-67140/2020\|EPI_ISL_651608\|202 | ···················· | ···················· | ···················· | ···················· | ···················· | ···················· |
| S | 8240 | hCoV-19/England/BIRM-67483/2020\|EPI_ISL_651610\|202 | ···················· | ···················· | ···················· | ···················· | ···················· | ···················· |
| S | 8241 | hCoV-19/England/BIRM-674CF/2020\|EPI_ISL_651611\|202 | ···················· | ···················· | ···················· | ···················· | ···················· | ···················· |
| S | 8242 | hCoV-19/England/BIRM-674DE/2020\|EPI_ISL_651612\|202 | ···················· | ···················· | ···················· | ···················· | ···················· | ···················· |
| S | 8244 | hCoV-19/England/BIRM-67526/2020\|EPI_ISL_651614\|202 | ···················· | ···················· | ···················· | ···················· | ···················· | ···················· |
| S | 8245 | hCoV-19/England/BIRM-67553/2020\|EPI_ISL_651615\|202 | ···················· | ·A·················· | ···················· | ···················· | ···················· | ···A················ |
| S | 8248 | hCoV-19/England/BIRM-675AE/2020\|EPI_ISL_651618\|202 | ···················· | ···················· | ···················· | ···················· | ···················· | ···················· |
| S | 8250 | hCoV-19/England/BIRM-67614/2020\|EPI_ISL_651620\|202 | ···················· | ···················· | ···················· | ···················· | ···················· | ···················· |
| S | 8251 | hCoV-19/England/BIRM-67623/2020\|EPI_ISL_651621\|202 | ···················· | ···················· | ···················· | ···················· | ···················· | ···················· |
| S | 8252 | hCoV-19/England/BIRM-676D8/2020\|EPI_ISL_651622\|202 | ···················· | ···················· | ···················· | ···················· | ···················· | ···················· |
| S | 8253 | hCoV-19/England/BIRM-6781E/2020\|EPI_ISL_651623\|202 | ···················· | ···················· | ···················· | ···················· | ···················· | ···················· |
| S | 8255 | hCoV-19/England/BIRM-67A27/2020\|EPI_ISL_651625\|202 | ···················· | ···················· | ···················· | ···················· | ···················· | ···················· |
| S | 8256 | hCoV-19/England/BIRM-67A45/2020\|EPI_ISL_651626\|202 | ···················· | ···················· | ···················· | ···················· | ···················· | ···················· |
| S | 8258 | hCoV-19/England/BIRM-67AEB/2020\|EPI_ISL_651628\|202 | ···················· | ···················· | ···················· | ···················· | ···················· | ···················· |
| S | 8260 | hCoV-19/England/BIRM-67C4F/2020\|EPI_ISL_651630\|202 | ···················· | ···················· | ···················· | ···················· | ···················· | ···················· |
| S | 8261 | hCoV-19/England/BIRM-67F19/2020\|EPI_ISL_651631\|202 | ···················· | ···················· | ···················· | ···················· | ···················· | ···················· |
| S | 8262 | hCoV-19/England/BIRM-67FCE/2020\|EPI_ISL_651632\|202 | ···················· | ···················· | ···················· | ···················· | ···················· | ···················· |
| S | 8263 | hCoV-19/England/BIRM-6A620/2020\|EPI_ISL_651633\|202 | ···················· | ···················· | ···················· | ···················· | ···················· | ···················· |
| S | 8264 | hCoV-19/England/BIRM-6A990/2020\|EPI_ISL_651634\|202 | ···················· | ···················· | ···················· | ···················· | ···················· | ···················· |
| S | 8265 | hCoV-19/England/BIRM-6A9AF/2020\|EPI_ISL_651635\|202 | ···················· | ···················· | ···················· | ···················· | ···················· | ···················· |
| S | 8266 | hCoV-19/England/BIRM-6A9CD/2020\|EPI_ISL_651636\|202 | ···················· | ···················· | ···················· | ···················· | ···················· | ···················· |
| S | 8267 | hCoV-19/England/BIRM-6A9FA/2020\|EPI_ISL_651637\|202 | ···················· | ···················· | ···················· | ···················· | ···················· | ···················· |
| S | 8268 | hCoV-19/England/BIRM-6AA24/2020\|EPI_ISL_651638\|202 | ···················· | ···················· | ···················· | ···················· | ···················· | ···················· |
| S | 8269 | hCoV-19/England/BIRM-6AA33/2020\|EPI_ISL_651639\|202 | ···················· | ···················· | ···················· | ···················· | ···················· | ···················· |
| S | 8270 | hCoV-19/England/BIRM-6AA60/2020\|EPI_ISL_651640\|202 | ···················· | ···················· | ···················· | ···················· | ···················· | ···················· |
| S | 8271 | hCoV-19/England/BIRM-6AA7F/2020\|EPI_ISL_651641\|202 | ···················· | ···················· | ···················· | ···················· | ···················· | ···················· |
| S | 8272 | hCoV-19/Scotland/QEUH-AACB65/2020\|EPI_ISL_652900\|2 | ···················· | ···················· | ···················· | ···················· | ···················· | ···················· |

| | | | | | | | | |
|---|---|---|---|---|---|---|---|---|
| S | 8273 | hCoV-19/Scotland/QEUH-AACB74/2020\|EPI_ISL_652901\|2 | .................... | .................... | .................... | .................... | .................... | .................... |
| S | 8274 | hCoV-19/Scotland/QEUH-AACB83/2020\|EPI_ISL_652902\|2 | .................... | .................... | .................... | .................... | .................... | .................... |
| S | 8275 | hCoV-19/Scotland/QEUH-AACBB0/2020\|EPI_ISL_652903\|2 | .................... | .................... | .................... | .................... | .................... | .................... |
| S | 8276 | hCoV-19/Scotland/QEUH-AACBCF/2020\|EPI_ISL_652904\|2 | .................... | .................... | .................... | .................... | .................... | .................... |
| S | 8277 | hCoV-19/Scotland/QEUH-AACBDE/2020\|EPI_ISL_652905\|2 | .................... | .................... | .................... | .................... | .................... | .................... |
| S | 8278 | hCoV-19/Scotland/QEUH-AACBFC/2020\|EPI_ISL_652906\|2 | .................... | .................... | .................... | .................... | .................... | .................... |
| S | 8279 | hCoV-19/Scotland/QEUH-AACC08/2020\|EPI_ISL_652907\|2 | .................... | .................... | .................... | .................... | .................... | .................... |
| S | 8280 | hCoV-19/Scotland/QEUH-AACC26/2020\|EPI_ISL_652908\|2 | .................... | .................... | .................... | .................... | .................... | .................... |
| S | 8281 | hCoV-19/England/QEUH-AACC80/2020\|EPI_ISL_652909\|20 | .................... | .................... | .................... | .................... | .................... | .................... |
| S | 8282 | hCoV-19/England/QEUH-AACC9F/2020\|EPI_ISL_652910\|20 | .................... | .................... | .................... | .................... | .................... | .................... |
| S | 8283 | hCoV-19/England/QEUH-AACCBD/2020\|EPI_ISL_652911\|20 | .................... | .................... | .................... | .................... | .................... | .................... |
| S | 8284 | hCoV-19/England/QEUH-AACCCC/2020\|EPI_ISL_652912\|20 | .................... | .................... | .................... | .................... | .................... | ..A................. |
| S | 8285 | hCoV-19/England/QEUH-AACD23/2020\|EPI_ISL_652913\|20 | .................... | .................... | .................... | .................... | .................... | .................... |
| S | 8286 | hCoV-19/Scotland/QEUH-AACD32/2020\|EPI_ISL_652914\|2 | .................... | .................... | .................... | .................... | .................... | .................... |
| S | 8287 | hCoV-19/England/QEUH-AACD9C/2020\|EPI_ISL_652915\|20 | .................... | .................... | .................... | .................... | .................... | .................... |
| S | 8288 | hCoV-19/Scotland/QEUH-AACDD8/2020\|EPI_ISL_652916\|2 | .................... | .................... | .................... | .................... | .................... | .................... |
| S | 8289 | hCoV-19/England/QEUH-AACDF6/2020\|EPI_ISL_652917\|20 | .................... | .................... | .................... | .................... | .................... | .................... |
| S | 8290 | hCoV-19/England/QEUH-AACE4E/2020\|EPI_ISL_652918\|20 | .................... | .................... | .................... | .................... | .................... | .................... |
| S | 8291 | hCoV-19/England/QEUH-AACE5D/2020\|EPI_ISL_652919\|20 | .................... | .................... | .................... | .................... | .................... | .................... |
| S | 8292 | hCoV-19/England/QEUH-AACE7B/2020\|EPI_ISL_652920\|20 | .................... | .................... | .................... | .................... | .................... | .................... |
| S | 8293 | hCoV-19/England/QEUH-AACEA8/2020\|EPI_ISL_652921\|20 | .................... | .................... | .................... | .................... | .................... | .................... |
| S | 8294 | hCoV-19/England/QEUH-AACED5/2020\|EPI_ISL_652922\|20 | .................... | .................... | .................... | .................... | .................... | .................... |
| S | 8295 | hCoV-19/England/QEUH-AACEE4/2020\|EPI_ISL_652923\|20 | .................... | .................... | .................... | .................... | .................... | .................... |
| S | 8296 | hCoV-19/England/QEUH-AACEF3/2020\|EPI_ISL_652924\|20 | .................... | .................... | .................... | .................... | .................... | .................... |
| S | 8297 | hCoV-19/England/QEUH-AACF0F/2020\|EPI_ISL_652925\|20 | .................... | .................... | .................... | .................... | .................... | .................... |
| S | 8298 | hCoV-19/England/QEUH-AACF1E/2020\|EPI_ISL_652926\|20 | .................... | .................... | .................... | .................... | .................... | .................... |
| S | 8299 | hCoV-19/England/QEUH-AACF3C/2020\|EPI_ISL_652927\|20 | .................... | .................... | .................... | .................... | .................... | .................... |
| S | 8300 | hCoV-19/England/QEUH-AACF4B/2020\|EPI_ISL_652928\|20 | .................... | .................... | .................... | .................... | ......T............. | .................... |
| S | 8301 | hCoV-19/England/QEUH-AACF5A/2020\|EPI_ISL_652929\|20 | .................... | .................... | .................... | .................... | .................... | .................... |
| S | 8302 | hCoV-19/England/QEUH-AACF69/2020\|EPI_ISL_652930\|20 | .................... | .................... | .................... | .................... | .................... | .................... |
| S | 8303 | hCoV-19/England/QEUH-AACFB4/2020\|EPI_ISL_652931\|20 | .................... | .................... | .................... | .................... | .................... | .................... |
| S | 8304 | hCoV-19/Scotland/QEUH-AACFC3/2020\|EPI_ISL_652932\|2 | .................... | .................... | .................... | .................... | .................... | .................... |
| S | 8305 | hCoV-19/England/QEUH-AAD00B/2020\|EPI_ISL_652933\|20 | .................... | .................... | .................... | .................... | .................... | .................... |
| S | 8306 | hCoV-19/England/QEUH-AAD047/2020\|EPI_ISL_652934\|20 | .................... | .................... | .................... | .................... | .................... | .................... |
| S | 8307 | hCoV-19/England/QEUH-AAD056/2020\|EPI_ISL_652935\|20 | .................... | .................... | .................... | .................... | .................... | .................... |
| S | 8308 | hCoV-19/England/QEUH-AAD083/2020\|EPI_ISL_652936\|20 | .................... | .................... | .................... | .................... | .................... | .................... |
| S | 8309 | hCoV-19/England/QEUH-AAD0A1/2020\|EPI_ISL_652937\|20 | .................... | .................... | .................... | .................... | .................... | .................... |
| S | 8310 | hCoV-19/England/QEUH-AAD0CF/2020\|EPI_ISL_652938\|20 | .................... | .................... | .................... | .................... | .................... | .................... |
| S | 8311 | hCoV-19/England/QEUH-AAD0DE/2020\|EPI_ISL_652939\|20 | .................... | .................... | .................... | .................... | .................... | .................... |
| S | 8312 | hCoV-19/Scotland/QEUH-AAD0ED/2020\|EPI_ISL_652940\|2 | .................... | .................... | .................... | .................... | .................... | .................... |
| S | 8313 | hCoV-19/England/QEUH-AAD117/2020\|EPI_ISL_652941\|20 | .................... | .................... | .................... | .................... | .................... | .................... |
| S | 8314 | hCoV-19/Scotland/QEUH-AAD126/2020\|EPI_ISL_652942\|2 | .................... | .................... | .................... | .................... | .................... | .................... |
| S | 8315 | hCoV-19/England/QEUH-AAD135/2020\|EPI_ISL_652943\|20 | .................... | .................... | .................... | .................... | .................... | .................... |
| S | 8316 | hCoV-19/England/QEUH-AAD144/2020\|EPI_ISL_652944\|20 | .................... | .................... | .................... | .................... | .................... | .................... |
| S | 8317 | hCoV-19/Scotland/QEUH-AAD153/2020\|EPI_ISL_652945\|2 | .................... | .................... | .................... | .................... | .................... | .................... |
| S | 8318 | hCoV-19/England/QEUH-AAD162/2020\|EPI_ISL_652946\|20 | .................... | .................... | .................... | .................... | .................... | .................... |
| S | 8319 | hCoV-19/England/QEUH-AADB28/2020\|EPI_ISL_652947\|20 | .................... | .................... | .................... | .................... | .................... | .................... |
| S | 8320 | hCoV-19/England/QEUH-AADB55/2020\|EPI_ISL_652948\|20 | .................... | .................... | .................... | .................... | .................... | .................... |
| S | 8321 | hCoV-19/England/QEUH-AADDC8/2020\|EPI_ISL_652949\|20 | .................... | .................... | .................... | .................... | .................... | .................... |
| S | 8322 | hCoV-19/England/QEUH-AADDD7/2020\|EPI_ISL_652950\|20 | .................... | .................... | .................... | .................... | .................... | .................... |
| S | 8323 | hCoV-19/England/QEUH-AADDE6/2020\|EPI_ISL_652951\|20 | .................... | .................... | .................... | .................... | .................... | .................... |
| S | 8324 | hCoV-19/Scotland/QEUH-AADDF5/2020\|EPI_ISL_652952\|2 | .................... | .................... | .................... | .................... | .................... | .................... |
| S | 8325 | hCoV-19/England/QEUH-AADE01/2020\|EPI_ISL_652953\|20 | .................... | .................... | .................... | .................... | .................... | .................... |
| S | 8326 | hCoV-19/Scotland/QEUH-AADE2F/2020\|EPI_ISL_652954\|2 | .................... | .................... | .................... | .................... | .................... | .................... |
| S | 8327 | hCoV-19/England/QEUH-AADE3E/2020\|EPI_ISL_652955\|20 | .................... | .................... | .................... | .................... | .................... | .................... |
| S | 8328 | hCoV-19/England/QEUH-AADE5C/2020\|EPI_ISL_652956\|20 | .................... | .................... | .................... | .................... | .................... | .................... |
| S | 8329 | hCoV-19/England/QEUH-AADE98/2020\|EPI_ISL_652957\|20 | .................... | .................... | .................... | .................... | .................... | .................... |
| S | 8330 | hCoV-19/England/QEUH-AADEB6/2020\|EPI_ISL_652958\|20 | .................... | .................... | .................... | .................... | .................... | .................... |
| S | 8331 | hCoV-19/England/QEUH-AADED4/2020\|EPI_ISL_652959\|20 | .................... | .................... | .................... | .................... | .................... | .................... |
| S | 8332 | hCoV-19/England/QEUH-AAE18F/2020\|EPI_ISL_652960\|20 | .................... | .................... | .................... | .................... | .................... | .................... |
| S | 8333 | hCoV-19/England/QEUH-AAE1AD/2020\|EPI_ISL_652961\|20 | .................... | .................... | .................... | .................... | .................... | .................... |
| S | 8334 | hCoV-19/England/QEUH-AAE1CB/2020\|EPI_ISL_652962\|20 | .................... | .................... | .................... | .................... | .................... | .................... |
| S | 8335 | hCoV-19/England/QEUH-AAE1DA/2020\|EPI_ISL_652963\|20 | .................... | .................... | .................... | .................... | ......T............. | .................... |
| S | 8336 | hCoV-19/England/QEUH-AAE1E9/2020\|EPI_ISL_652964\|20 | .................... | .................... | .................... | .................... | ....T............... | .................... |
| S | 8337 | hCoV-19/England/QEUH-AAE25F/2020\|EPI_ISL_652965\|20 | .................... | .................... | .................... | .................... | .................... | .................... |
| S | 8338 | hCoV-19/Scotland/QEUH-AAE2C8/2020\|EPI_ISL_652966\|2 | .................... | .................... | .................... | .................... | .................... | .................... |
| S | 8339 | hCoV-19/Scotland/QEUH-AAE2D7/2020\|EPI_ISL_652967\|2 | .................... | .................... | .................... | .................... | .................... | .................... |
| S | 8340 | hCoV-19/Scotland/QEUH-AAE2F5/2020\|EPI_ISL_652968\|2 | .................... | .................... | .................... | .................... | .................... | .................... |
| S | 8341 | hCoV-19/Scotland/QEUH-AAE301/2020\|EPI_ISL_652969\|2 | .................... | .................... | .................... | .................... | .................... | .................... |
| S | 8342 | hCoV-19/Scotland/QEUH-AAE37A/2020\|EPI_ISL_652970\|2 | .................... | .................... | .................... | .................... | .................... | .................... |
| S | 8343 | hCoV-19/Scotland/QEUH-AAE398/2020\|EPI_ISL_652971\|2 | .................... | .................... | .................... | .................... | .................... | .................... |
| S | 8344 | hCoV-19/Scotland/QEUH-AAE3B6/2020\|EPI_ISL_652972\|2 | .................... | .................... | .................... | .................... | .................... | .................... |
| S | 8345 | hCoV-19/England/QEUH-B6BD6B/2020\|EPI_ISL_655170\|20 | .................... | .................... | .................... | .................... | .................... | .................... |
| S | 8346 | hCoV-19/England/QEUH-B68CF8/2020\|EPI_ISL_655171\|20 | .................... | .................... | .................... | .................... | .................... | .................... |
| S | 8347 | hCoV-19/England/QEUH-B6C19B/2020\|EPI_ISL_655172\|20 | .................... | .................... | .................... | .................... | .................... | ..A................. |
| S | 8348 | hCoV-19/England/QEUH-B6C8B4/2020\|EPI_ISL_655173\|20 | .................... | .................... | .................... | .................... | .................... | .................... |
| S | 8349 | hCoV-19/England/QEUH-B6BDB6/2020\|EPI_ISL_655174\|20 | .................... | .................... | .................... | .................... | ......T............. | .................... |
| S | 8350 | hCoV-19/England/QEUH-B6C1AA/2020\|EPI_ISL_655175\|20 | .................... | .................... | .................... | .................... | .................... | .................... |
| S | 8351 | hCoV-19/England/QEUH-B68812/2020\|EPI_ISL_655176\|20 | .................... | .................... | .................... | .................... | .................... | .................... |
| S | 8352 | hCoV-19/England/QEUH-B67CCC/2020\|EPI_ISL_655178\|20 | .................... | .................... | .................... | .................... | .................... | .................... |

| | | | | | | | |
|---|---|---|---|---|---|---|---|
| S | 8353 hCoV-19/England/QEUH-B67CF9/2020\|EPI_ISL_655179\|20 | ··················· | ··················· | ···················· | ··················· | ··················· | ··················· |
| S | 8354 hCoV-19/England/QEUH-B68A85/2020\|EPI_ISL_655180\|20 | ··················· | ··················· | ···················· | ··················· | ··················· | ··················· |
| S | 8355 hCoV-19/England/QEUH-B69C24/2020\|EPI_ISL_655182\|20 | ··················· | ··················· | ···················· | ··················· | ··················· | ··················· |
| S | 8356 hCoV-19/England/QEUH-B685DF/2020\|EPI_ISL_655183\|20 | ··················· | ··················· | ···················· | ··················· | ··················· | ··················· |
| S | 8357 hCoV-19/England/QEUH-B69EF1/2020\|EPI_ISL_655185\|20 | ··················· | ··················· | ···················· | ··················· | ··················· | ··················· |
| S | 8358 hCoV-19/England/QEUH-B69B90/2020\|EPI_ISL_655188\|20 | ··················· | ··················· | ···················· | ··················· | ··················· | ··················· |
| S | 8359 hCoV-19/England/QEUH-B6BE59/2020\|EPI_ISL_655189\|20 | ··················· | ··················· | ···················· | ··················· | ··················· | ··················· |
| S | 8360 hCoV-19/England/QEUH-B6ACBA/2020\|EPI_ISL_655190\|20 | ··················· | ··················· | ···················· | ··················· | ··················· | ··················· |
| S | 8361 hCoV-19/England/QEUH-B6C34A/2020\|EPI_ISL_655191\|20 | ··················· | ··················· | ···················· | ··················· | ··················· | ··················· |
| S | 8362 hCoV-19/England/QEUH-B6884F/2020\|EPI_ISL_655192\|20 | ··················· | ··················· | ···················· | ··················· | ··················· | ··················· |
| S | 8363 hCoV-19/England/QEUH-B69F76/2020\|EPI_ISL_655193\|20 | ··················· | ··················· | ···················· | ··················· | ··················· | ··················· |
| S | 8364 hCoV-19/England/QEUH-B68CDA/2020\|EPI_ISL_655194\|20 | ··················· | ··················· | ···················· | ··················· | ··················· | ··················· |
| S | 8365 hCoV-19/England/QEUH-B6AD02/2020\|EPI_ISL_655196\|20 | ··················· | ··················· | ···················· | ··················· | ··················· | ··················· |
| S | 8366 hCoV-19/England/QEUH-B69BAF/2020\|EPI_ISL_655197\|20 | ··················· | ··················· | ···················· | ··················· | ··················· | ··················· |
| S | 8367 hCoV-19/England/QEUH-B695A1/2020\|EPI_ISL_655198\|20 | ··················· | ··················· | ···················· | ··················· | ··················· | ··················· |
| S | 8368 hCoV-19/England/QEUH-B68539/2020\|EPI_ISL_655199\|20 | ··················· | ··················· | ···················· | ··················· | ··················· | ··················· |
| S | 8369 hCoV-19/England/QEUH-B6AD3F/2020\|EPI_ISL_655206\|20 | ··················· | ··················· | ···················· | ··················· | ··················· | ··············A···· |
| S | 8370 hCoV-19/England/QEUH-B6B958/2020\|EPI_ISL_655207\|20 | ··················· | ··················· | ···················· | ··················· | ··················· | ··················· |
| S | 8371 hCoV-19/England/QEUH-B6B994/2020\|EPI_ISL_655208\|20 | ··················· | ··················· | ···················· | ··················· | ··················· | ··················· |
| S | 8372 hCoV-19/England/QEUH-B685EE/2020\|EPI_ISL_655209\|20 | ··················· | ··················· | ···················· | ··················· | ··················· | ··················· |
| S | 8373 hCoV-19/England/QEUH-B68830/2020\|EPI_ISL_655210\|20 | ··················· | ··················· | ···················· | ··················· | ··················· | ··················· |
| S | 8374 hCoV-19/England/QEUH-B6A2B8/2020\|EPI_ISL_655211\|20 | ··················· | ··················· | ···················· | ··················· | ··················· | ··················· |
| S | 8375 hCoV-19/England/QEUH-B6B93A/2020\|EPI_ISL_655214\|20 | ··················· | ··················· | ···················· | ··················· | ··················· | ··················· |
| S | 8376 hCoV-19/England/QEUH-B68821/2020\|EPI_ISL_655216\|20 | ··················· | ··················· | ···················· | ··················· | ··················· | ··················· |
| S | 8377 hCoV-19/England/QEUH-B6AC9C/2020\|EPI_ISL_655217\|20 | ··················· | ··················· | ···················· | ··················· | ··················· | ··················· |
| S | 8378 hCoV-19/England/QEUH-B6950B/2020\|EPI_ISL_655219\|20 | ··················· | ··················· | ···················· | ··················· | ··················· | ··················· |
| S | 8379 hCoV-19/England/QEUH-B6B985/2020\|EPI_ISL_655220\|20 | ··················· | ··················· | ···················· | ··················· | ··················· | ··················· |
| S | 8380 hCoV-19/England/QEUH-B6C0AD/2020\|EPI_ISL_655221\|20 | ··················· | ··················· | ···················· | ··················· | ··················· | ··················· |
| S | 8381 hCoV-19/England/QEUH-B6BA46/2020\|EPI_ISL_655222\|20 | ··················· | ··················· | ···················· | ··················· | ··················· | ··················· |
| S | 8382 hCoV-19/England/QEUH-B6BDD4/2020\|EPI_ISL_655223\|20 | ··················· | ··················· | ···················· | ··················· | ··················· | ··················· |
| S | 8383 hCoV-19/England/QEUH-B69BFA/2020\|EPI_ISL_655226\|20 | ··················· | ··················· | ···················· | ··················· | ··················· | ··················· |
| S | 8384 hCoV-19/England/QEUH-B6C025/2020\|EPI_ISL_655227\|20 | ··················· | ··················· | ···················· | ··················· | ··················· | ··················· |
| S | 8385 hCoV-19/England/QEUH-B68AA3/2020\|EPI_ISL_655239\|20 | ··················· | ··················· | ···················· | ··················· | ··················· | ··················· |
| S | 8386 hCoV-19/England/QEUH-B66A5A/2020\|EPI_ISL_655242\|20 | ··················· | ··················· | ···················· | ··················· | ··················· | ··················· |
| S | 8387 hCoV-19/England/QEUH-B67D6F/2020\|EPI_ISL_655243\|20 | ··················· | ··················· | ···················· | ··················· | ··················· | ··················· |
| S | 8388 hCoV-19/England/QEUH-B69547/2020\|EPI_ISL_655244\|20 | ··················· | ··················· | ···················· | ··················· | ··················· | ··················· |
| S | 8389 hCoV-19/England/QEUH-B68C8F/2020\|EPI_ISL_655247\|20 | ··················· | ·A················· | ···················· | ··················· | ··················· | ··················· |
| S | 8390 hCoV-19/England/ALDP-B62405/2020\|EPI_ISL_656538\|20 | ··················· | ··················· | ···················· | ··················· | ··················· | ··················· |
| S | 8391 hCoV-19/England/ALDP-B639BA/2020\|EPI_ISL_656539\|20 | ··················· | ··················· | ···················· | ··················· | ··················· | ··················· |
| S | 8392 hCoV-19/England/ALDP-B6248D/2020\|EPI_ISL_656540\|20 | ··················· | ··················· | ···················· | ··················· | ··················· | ··················· |
| S | 8393 hCoV-19/England/ALDP-B62881/2020\|EPI_ISL_656541\|20 | ··················· | ··················· | ···················· | ··················· | ··················· | ··················· |
| S | 8394 hCoV-19/England/ALDP-B6210E/2020\|EPI_ISL_656542\|20 | ··················· | ··················· | ···················· | ··················· | ··················· | ··················· |
| S | 8395 hCoV-19/England/ALDP-B632DD/2020\|EPI_ISL_656543\|20 | ··················· | ··················· | ···················· | ··················· | ··················· | ··················· |
| S | 8396 hCoV-19/England/ALDP-B63BF0/2020\|EPI_ISL_656547\|20 | ··················· | ··················· | ···················· | ··················· | ··················· | ··················· |
| S | 8397 hCoV-19/England/ALDP-B633DA/2020\|EPI_ISL_656549\|20 | ··················· | ··················· | ···················· | ··················· | ··················· | ··················· |
| S | 8398 hCoV-19/England/ALDP-B63B2D/2020\|EPI_ISL_656550\|20 | ··················· | ··················· | ···················· | ··················· | ··················· | ··················· |
| S | 8399 hCoV-19/England/ALDP-B62511/2020\|EPI_ISL_656551\|20 | ··················· | ·A················· | ···················· | ··················· | ··················· | ··A················ |
| S | 8400 hCoV-19/England/ALDP-B636A4/2020\|EPI_ISL_656552\|20 | ··················· | ··················· | ···················· | ··················· | ··················· | ··················· |
| S | 8401 hCoV-19/England/ALDP-B63835/2020\|EPI_ISL_656554\|20 | ··················· | ··················· | ···················· | ··················· | ··················· | ··················· |
| S | 8402 hCoV-19/England/ALDP-B63A4E/2020\|EPI_ISL_656555\|20 | ··················· | ··················· | ···················· | ··················· | ··················· | ··················· |
| S | 8403 hCoV-19/England/ALDP-B62A9A/2020\|EPI_ISL_656556\|20 | ··················· | ········T·········· | ···················· | ··················· | ··················· | ··················· |
| S | 8404 hCoV-19/England/ALDP-B6212C/2020\|EPI_ISL_656557\|20 | ··················· | ··················· | ···················· | ··················· | ··················· | ··················· |
| S | 8405 hCoV-19/England/ALDP-B624C9/2020\|EPI_ISL_656558\|20 | ··················· | ··················· | ···················· | ··················· | ··················· | ··················· |
| S | 8406 hCoV-19/England/ALDP-B6357A/2020\|EPI_ISL_656573\|20 | ··················· | ··················· | ···················· | ··················· | ··················· | ··················· |
| S | 8407 hCoV-19/England/ALDP-B63AA8/2020\|EPI_ISL_656574\|20 | ··················· | ··················· | ···················· | ··················· | ··················· | ··················· |
| S | 8408 hCoV-19/England/ALDP-B620A7/2020\|EPI_ISL_656575\|20 | ··················· | ··················· | ···················· | ··················· | ··················· | ··················· |
| S | 8409 hCoV-19/England/ALDP-B62274/2020\|EPI_ISL_656576\|20 | ··················· | ··················· | ···················· | ··················· | ··················· | ··················· |
| S | 8410 hCoV-19/England/ALDP-B63880/2020\|EPI_ISL_656577\|20 | ··················· | ··················· | ···················· | ··················· | ··················· | ··················· |
| S | 8411 hCoV-19/England/ALDP-B631D0/2020\|EPI_ISL_656579\|20 | ··················· | ··················· | ···················· | ··················· | ··················· | ··················· |
| S | 8412 hCoV-19/England/CAMC-B5208C/2020\|EPI_ISL_657868\|20 | ··················· | ··················· | ···················· | ··················· | ··················· | ··················· |
| S | 8413 hCoV-19/England/MILK-B54CD1/2020\|EPI_ISL_657870\|20 | ··················· | ··················· | ···················· | ··················· | ··················· | ··················· |
| S | 8414 hCoV-19/England/CAMC-B51506/2020\|EPI_ISL_657871\|20 | ··················· | ··················· | ···················· | ··················· | ··················· | ··················· |
| S | 8415 hCoV-19/England/MILK-B54F6F/2020\|EPI_ISL_657872\|20 | ··················· | ··················· | ···················· | ··················· | ··················· | ··················· |
| S | 8416 hCoV-19/England/MILK-B5557B/2020\|EPI_ISL_657874\|20 | ··················· | ··················· | ···················· | ··················· | ··················· | ··················· |
| S | 8417 hCoV-19/England/CAMC-B51F17/2020\|EPI_ISL_657875\|20 | ··················· | ··················· | ···················· | ··················· | ··················· | ··················· |
| S | 8418 hCoV-19/England/CAMC-B510BA/2020\|EPI_ISL_657876\|20 | ··················· | ··················· | ···················· | ··················· | ··················· | ··················· |
| S | 8419 hCoV-19/England/MILK-B55414/2020\|EPI_ISL_657879\|20 | ··················· | ··················· | ···················· | ··················· | ··················· | ··················· |
| S | 8420 hCoV-19/England/CAMC-B511D5/2020\|EPI_ISL_657881\|20 | ··················· | ··················· | ···················· | ··················· | ··················· | ··················· |
| S | 8421 hCoV-19/England/CAMC-B51C6B/2020\|EPI_ISL_657883\|20 | ··················· | ··················· | ···················· | ··················· | ··················· | ··················· |
| S | 8422 hCoV-19/England/CAMC-B51524/2020\|EPI_ISL_657884\|20 | ··················· | ··················· | ···················· | ··················· | ··················· | ··················· |
| S | 8423 hCoV-19/England/CAMC-B510AB/2020\|EPI_ISL_657885\|20 | ··················· | ··················· | ···················· | ··················· | ··················· | ··················· |
| S | 8424 hCoV-19/England/MILK-B54F8D/2020\|EPI_ISL_657886\|20 | ··················· | ··················· | ···················· | ··················· | ··················· | ··················· |
| S | 8425 hCoV-19/England/MILK-B55089/2020\|EPI_ISL_657887\|20 | ··················· | ··················· | ···················· | ··················· | ··················· | ··················· |
| S | 8426 hCoV-19/England/CAMC-B51EB0/2020\|EPI_ISL_657888\|20 | ·Y················· | ··················· | ···················· | ··················· | ··················· | ··················· |
| S | 8427 hCoV-19/England/MILK-B552DE/2020\|EPI_ISL_657889\|20 | ··················· | ··················· | ···················· | ··················· | ··················· | ··················· |
| S | 8428 hCoV-19/USA/WI-GMF-30065/2020\|EPI_ISL_661025\|2020- | ··················· | ··················· | ···················· | ··················· | ··················· | ··················· |
| S | 8429 hCoV-19/USA/WI-GMF-30076/2020\|EPI_ISL_661026\|2020- | ··················· | ··················· | ···················· | ··················· | ··················· | ··················· |
| S | 8430 hCoV-19/USA/WI-GMF-30188/2020\|EPI_ISL_661027\|2020- | ··················· | ··················· | ···················· | ··················· | ··················· | ··················· |
| S | 8431 hCoV-19/USA/WI-GMF-30231/2020\|EPI_ISL_661028\|2020- | ··················· | ··················· | ···················· | ··················· | ··················· | ··················· |
| S | 8432 hCoV-19/USA/WI-GMF-30246/2020\|EPI_ISL_661029\|2020- | ··················· | ··················· | ···················· | ··················· | ··················· | ··················· |

| | | | | | | | | |
|---|---|---|---|---|---|---|---|---|
| S | 8433 | hCoV-19/USA/WI-GMF-30249/2020\|EPI_ISL_661030\|2020- | .................... | .A.................. | ..................... | .................... | .................... | .................... |
| S | 8434 | hCoV-19/USA/WI-GMF-30272/2020\|EPI_ISL_661031\|2020- | .................... | .................... | ..................... | .................... | .................... | .................... |
| S | 8435 | hCoV-19/USA/WI-GMF-30278/2020\|EPI_ISL_661032\|2020- | .................... | .................... | ..................... | .................... | .................... | .................... |
| S | 8436 | hCoV-19/USA/WI-GMF-30352/2020\|EPI_ISL_661033\|2020- | .................... | ..C................. | ..................... | .................... | .................... | .................... |
| S | 8437 | hCoV-19/USA/WI-GMF-30445/2020\|EPI_ISL_661034\|2020- | .................... | ..C................. | ..................... | .................... | .................... | .................... |
| S | 8438 | hCoV-19/USA/WI-GMF-30475/2020\|EPI_ISL_661036\|2020- | .................... | .................... | ..................... | .................... | .................... | .................... |
| S | 8439 | hCoV-19/USA/WI-GMF-30476/2020\|EPI_ISL_661037\|2020- | .................... | .................... | ..................... | .................... | .................... | .................... |
| S | 8440 | hCoV-19/USA/WI-GMF-30528/2020\|EPI_ISL_661038\|2020- | .................... | .................... | ..................... | .................... | .................... | .................... |
| S | 8441 | hCoV-19/USA/WI-GMF-30606/2020\|EPI_ISL_661039\|2020- | .................... | .................... | ..................... | .................... | .................... | .................... |
| S | 8442 | hCoV-19/USA/WI-GMF-30619/2020\|EPI_ISL_661040\|2020- | .................... | .................... | ..................... | .................... | .................... | .................... |
| S | 8443 | hCoV-19/USA/WI-GMF-30624/2020\|EPI_ISL_661041\|2020- | .................... | .................... | ..................... | .................... | .................... | .................... |
| S | 8444 | hCoV-19/USA/WI-GMF-30626/2020\|EPI_ISL_661042\|2020- | .................... | .................... | ..................... | .................... | .................... | .................... |
| S | 8445 | hCoV-19/USA/WI-GMF-30636/2020\|EPI_ISL_661043\|2020- | .................... | .................... | ..................... | .................... | .................... | .................... |
| S | 8446 | hCoV-19/USA/WI-GMF-30639/2020\|EPI_ISL_661044\|2020- | .................... | .................... | ..................... | .................... | .................... | .................... |
| S | 8447 | hCoV-19/USA/WI-GMF-30662/2020\|EPI_ISL_661045\|2020- | .................... | .................... | ..................... | .................... | .................... | .................... |
| S | 8448 | hCoV-19/USA/WI-GMF-30678/2020\|EPI_ISL_661046\|2020- | .................... | .................... | ..................... | .................... | .................... | .................... |
| S | 8449 | hCoV-19/England/ALDP-B48274/2020\|EPI_ISL_649290\|20 | .................... | .................... | ..................... | .................... | .................... | .................... |
| S | 8450 | hCoV-19/England/ALDP-B484D8/2020\|EPI_ISL_649291\|20 | .................... | .................... | ..................... | .................... | .................... | .................... |
| S | 8451 | hCoV-19/England/ALDP-B47DDE/2020\|EPI_ISL_649293\|20 | .................... | .................... | ..................... | .................... | .................... | .................... |
| S | 8452 | hCoV-19/England/ALDP-B48362/2020\|EPI_ISL_649294\|20 | .................... | .................... | ..................... | .................... | .................... | .................... |
| S | 8453 | hCoV-19/England/ALDP-B484AB/2020\|EPI_ISL_649295\|20 | .................... | .................... | ..................... | .................... | .................... | .................... |
| S | 8454 | hCoV-19/England/ALDP-B484F6/2020\|EPI_ISL_649296\|20 | .................... | .................... | ..................... | .................... | .................... | .................... |
| S | 8455 | hCoV-19/England/ALDP-B48168/2020\|EPI_ISL_649297\|20 | .................... | .................... | ..................... | .................... | .................... | .................... |
| S | 8456 | hCoV-19/England/ALDP-B4857B/2020\|EPI_ISL_649298\|20 | .................... | .................... | ..................... | .................... | .................... | .................... |
| S | 8457 | hCoV-19/England/ALDP-B4858A/2020\|EPI_ISL_649299\|20 | .................... | .................... | ..................... | .................... | .................... | .................... |
| S | 8458 | hCoV-19/England/ALDP-B4846F/2020\|EPI_ISL_649330\|20 | .................... | .................... | ..................... | .................... | .................... | .................... |
| S | 8459 | hCoV-19/England/ALDP-B490A6/2020\|EPI_ISL_649331\|20 | .................... | .................... | ..................... | .................... | .................... | .................... |
| S | 8460 | hCoV-19/England/ALDP-B48256/2020\|EPI_ISL_649332\|20 | .................... | .................... | ..................... | .................... | .................... | .................... |
| S | 8461 | hCoV-19/England/ALDP-B481C2/2020\|EPI_ISL_649333\|20 | .................... | .................... | ..................... | .................... | .................... | .................... |
| S | 8462 | hCoV-19/England/ALDP-B481E0/2020\|EPI_ISL_649335\|20 | .................... | .................... | ..................... | .................... | .................... | .................... |
| S | 8463 | hCoV-19/England/ALDP-B48441/2020\|EPI_ISL_649338\|20 | .................... | .................... | ..................... | .................... | .................... | .................... |
| S | 8464 | hCoV-19/England/ALDP-B48836/2020\|EPI_ISL_649339\|20 | .................... | .................... | ..................... | .................... | .................... | .................... |
| S | 8465 | hCoV-19/England/ALDP-B482ED/2020\|EPI_ISL_649341\|20 | .................... | .................... | ..................... | .................... | .................... | .................... |
| S | 8466 | hCoV-19/England/ALDP-B47E44/2020\|EPI_ISL_649342\|20 | .................... | .................... | ..................... | .................... | .................... | .................... |
| S | 8467 | hCoV-19/England/ALDP-B4820B/2020\|EPI_ISL_649343\|20 | .................... | .................... | ..................... | .................... | .................... | .................... |
| S | 8468 | hCoV-19/England/ALDP-B481B3/2020\|EPI_ISL_649344\|20 | .................... | .................... | ..................... | .................... | .................... | .................... |
| S | 8469 | hCoV-19/England/ALDP-B47EBD/2020\|EPI_ISL_649345\|20 | .................... | .................... | ..................... | .................... | .................... | .................... |
| S | 8470 | hCoV-19/England/ALDP-B48414/2020\|EPI_ISL_649347\|20 | .................... | .................... | ..................... | .................... | .................... | .................... |
| S | 8471 | hCoV-19/England/ALDP-B48DA0/2020\|EPI_ISL_649348\|20 | .................... | .................... | ..................... | .................... | .................... | .................... |
| S | 8472 | hCoV-19/England/ALDP-B4862D/2020\|EPI_ISL_649349\|20 | .................... | .................... | ..................... | .................... | .................... | .................... |
| S | 8473 | hCoV-19/England/ALDP-B49440/2020\|EPI_ISL_649350\|20 | .................... | .................... | ..................... | .................... | .................... | .................... |
| S | 8474 | hCoV-19/England/ALDP-B481FF/2020\|EPI_ISL_649351\|20 | .................... | .................... | ..................... | .................... | .................... | .................... |
| S | 8475 | hCoV-19/England/ALDP-B48159/2020\|EPI_ISL_649352\|20 | .................... | .................... | ..................... | .................... | .................... | .................... |
| S | 8476 | hCoV-19/England/ALDP-B50EFD/2020\|EPI_ISL_649354\|20 | .................... | .................... | ..................... | .................... | .................... | .................... |
| S | 8477 | hCoV-19/England/ALDP-B4854E/2020\|EPI_ISL_649355\|20 | .................... | .................... | ..................... | .................... | .................... | .................... |
| S | 8478 | hCoV-19/England/ALDP-B4906A/2020\|EPI_ISL_649357\|20 | .................... | .................... | ..................... | .................... | .................... | .................... |
| S | 8479 | hCoV-19/England/ALDP-B4806B/2020\|EPI_ISL_649358\|20 | .................... | .................... | ..................... | .................... | .................... | .................... |
| S | 8480 | hCoV-19/Scotland/ALDP-B48450/2020\|EPI_ISL_649359\|2 | .................... | .................... | ..................... | .................... | .................... | .................... |
| S | 8481 | hCoV-19/England/ALDP-B47F32/2020\|EPI_ISL_649360\|20 | .................... | .................... | ..................... | .................... | .................... | .................... |
| S | 8482 | hCoV-19/England/ALDP-B491D0/2020\|EPI_ISL_649361\|20 | .................... | .................... | ..................... | .................... | ......T............. | .................... |
| S | 8483 | hCoV-19/England/MILK-B504DD/2020\|EPI_ISL_649362\|20 | .................... | .................... | ..................... | .................... | .................... | .................... |
| S | 8484 | hCoV-19/England/ALDP-B47EEA/2020\|EPI_ISL_649363\|20 | .................... | .................... | ..................... | .................... | .................... | .................... |
| S | 8485 | hCoV-19/England/ALDP-B48933/2020\|EPI_ISL_649364\|20 | .................... | .................... | ..................... | .................... | .................... | .................... |
| S | 8486 | hCoV-19/England/ALDP-B47ECC/2020\|EPI_ISL_649365\|20 | .................... | .................... | ..................... | .................... | .................... | .................... |
| S | 8487 | hCoV-19/England/ALDP-B4910D/2020\|EPI_ISL_649366\|20 | .................... | .................... | ..................... | .................... | .................... | .................... |
| S | 8488 | hCoV-19/England/ALDP-B47E17/2020\|EPI_ISL_649367\|20 | .................... | .................... | ..................... | .................... | .................... | .................... |
| S | 8489 | hCoV-19/England/ALDP-B4FB09/2020\|EPI_ISL_649368\|20 | .................... | .................... | ..................... | .................... | .................... | .................... |
| S | 8490 | hCoV-19/England/ALDP-B50060/2020\|EPI_ISL_649369\|20 | .................... | .................... | ..................... | .................... | .................... | .................... |
| S | 8491 | hCoV-19/England/ALDP-B505DA/2020\|EPI_ISL_649370\|20 | .................... | .................... | ..................... | .................... | .................... | .................... |
| S | 8492 | hCoV-19/England/ALDP-B485F3/2020\|EPI_ISL_649372\|20 | .................... | .................... | ..................... | .................... | .................... | .................... |
| S | 8493 | hCoV-19/England/ALDP-B491B2/2020\|EPI_ISL_649373\|20 | .................... | .................... | ..................... | .................... | .................... | .................... |
| S | 8494 | hCoV-19/England/ALDP-B485E4/2020\|EPI_ISL_649374\|20 | .................... | .................... | ..................... | .................... | .................... | .................... |
| S | 8495 | hCoV-19/England/ALDP-B49194/2020\|EPI_ISL_649375\|20 | .................... | .................... | ..................... | .................... | .................... | .................... |
| S | 8496 | hCoV-19/England/ALDP-B47DA1/2020\|EPI_ISL_649376\|20 | .................... | .................... | ..................... | .................... | .................... | .................... |
| S | 8497 | hCoV-19/England/ALDP-B48502/2020\|EPI_ISL_649377\|20 | .................... | .................... | ..................... | .................... | .................... | .................... |
| S | 8498 | hCoV-19/England/ALDP-B47FD8/2020\|EPI_ISL_649378\|20 | .................... | .................... | ..................... | .................... | .................... | .................... |
| S | 8499 | hCoV-19/England/ALDP-B49246/2020\|EPI_ISL_649379\|20 | .................... | .................... | ..................... | .................... | .................... | .................... |
| S | 8500 | hCoV-19/England/ALDP-B48687/2020\|EPI_ISL_649381\|20 | .................... | .................... | ..................... | .................... | .................... | .................... |
| S | 8501 | hCoV-19/England/ALDP-B47FC9/2020\|EPI_ISL_649382\|20 | .................... | .................... | ..................... | .................... | .................... | .................... |
| S | 8502 | hCoV-19/England/ALDP-B47E35/2020\|EPI_ISL_649383\|20 | .................... | .................... | ..................... | .................... | .................... | .................... |
| S | 8503 | hCoV-19/England/ALDP-B480F2/2020\|EPI_ISL_649384\|20 | .................... | .................... | ..................... | .................... | .................... | .................... |
| S | 8504 | hCoV-19/England/ALDP-B47EF9/2020\|EPI_ISL_649385\|20 | .................... | .................... | ..................... | .................... | .................... | .................... |
| S | 8505 | hCoV-19/England/MILK-B50534/2020\|EPI_ISL_649386\|20 | .................... | .A.................. | ..................... | .................... | .................... | ..A................. |
| S | 8506 | hCoV-19/England/ALDP-B48DFB/2020\|EPI_ISL_649387\|20 | .................... | .................... | ..................... | .................... | .................... | .................... |
| S | 8507 | hCoV-19/England/ALDP-B4F6EA/2020\|EPI_ISL_649389\|20 | .................... | .................... | ..................... | .................... | .................... | .................... |
| S | 8508 | hCoV-19/England/BIRM-6AA9D/2020\|EPI_ISL_651642\|202 | .................... | .................... | ..................... | .................... | .................... | .................... |
| S | 8509 | hCoV-19/England/BIRM-6AAAC/2020\|EPI_ISL_651643\|202 | .................... | .................... | ..................... | .................... | .................... | .................... |
| S | 8510 | hCoV-19/England/BIRM-6AABB/2020\|EPI_ISL_651644\|202 | .................... | .................... | ..................... | .................... | .................... | .................... |
| S | 8511 | hCoV-19/England/BIRM-6AACA/2020\|EPI_ISL_651645\|202 | .................... | .................... | ..................... | .................... | .................... | .................... |
| S | 8512 | hCoV-19/England/BIRM-6AAD9/2020\|EPI_ISL_651646\|202 | .................... | .................... | ..................... | .................... | .................... | .................... |

```
S    8513 hCoV-19/England/BIRM-6AAE8/2020|EPI_ISL_651647|202    ····················  ····················  ·····················  ····················  ····················  ····················
S    8514 hCoV-19/England/BIRM-6AAF7/2020|EPI_ISL_651648|202    ····················  ····················  ·····················  ····················  ····················  ····················
S    8515 hCoV-19/England/BIRM-6AB30/2020|EPI_ISL_651649|202    ····················  ····················  ·····················  ····················  ····················  ····················
S    8517 hCoV-19/England/BIRM-6AB6D/2020|EPI_ISL_651651|202    ····················  ····················  ·····················  ····················  ····················  ····················
S    8518 hCoV-19/England/BIRM-6ABB8/2020|EPI_ISL_651652|202    ····················  ····················  ·····················  ····················  ····················  ····················
S    8519 hCoV-19/England/CAMB-1B85DA/2020|EPI_ISL_651653|20    ····················  ····················  ·····················  ····················  ····················  ····················
S    8520 hCoV-19/England/CAMB-1B8640/2020|EPI_ISL_651654|20    ····················  ····················  ·····················  ····················  ····················  ····················
S    8521 hCoV-19/England/CAMB-1B867D/2020|EPI_ISL_651655|20    ····················  ····················  ·····················  ····················  ····················  ····················
S    8522 hCoV-19/England/CAMB-1B8701/2020|EPI_ISL_651656|20    ····················  ····················  ·····················  ····················  ······T·············  ····················
S    8523 hCoV-19/England/CAMB-1B877A/2020|EPI_ISL_651657|20    ····················  ····················  ·····················  ····················  ····················  ····················
S    8524 hCoV-19/England/CAMB-1B8798/2020|EPI_ISL_651658|20    ····················  ····················  ·····················  ····················  ····················  ····················
S    8525 hCoV-19/England/CAMB-1B87C5/2020|EPI_ISL_651659|20    ····················  ····················  ·····················  ····················  ····················  ····················
S    8526 hCoV-19/England/CAMB-1B87E3/2020|EPI_ISL_651660|20    ····················  ·A··················  ·····················  ····················  ····················  ···A················
S    8527 hCoV-19/England/CAMB-1B87F2/2020|EPI_ISL_651661|20    ····················  ····················  ·····················  ····················  ····················  ····················
S    8528 hCoV-19/England/CAMB-1B881D/2020|EPI_ISL_651662|20    ····················  ····················  ·····················  ····················  ····················  ····················
S    8530 hCoV-19/England/CAMB-1B884A/2020|EPI_ISL_651664|20    ····················  ····················  ·····················  ····················  ····················  ····················
S    8532 hCoV-19/England/CAMB-1B88B3/2020|EPI_ISL_651666|20    ····················  ····················  ·····················  ····················  ····················  ····················
S    8533 hCoV-19/England/CAMB-1B88FF/2020|EPI_ISL_651667|20    ····················  ····················  ·····················  ····················  ····················  ····················
S    8534 hCoV-19/England/CAMB-1B890B/2020|EPI_ISL_651668|20    ····················  ·A··················  ·····················  ····················  ····················  ····················
S    8535 hCoV-19/England/CAMB-1B891A/2020|EPI_ISL_651669|20    ····················  ····················  ·····················  ····················  ····················  ····················
S    8536 hCoV-19/England/CAMB-1B8938/2020|EPI_ISL_651670|20    ····················  ····················  ·····················  ····················  ····················  ····················
S    8537 hCoV-19/England/CAMB-1B8947/2020|EPI_ISL_651671|20    ····················  ····················  ·····················  ····················  ····················  ····················
S    8538 hCoV-19/England/CAMB-1B8956/2020|EPI_ISL_651672|20    ····················  ····················  ·····················  ····················  ····················  ····················
S    8539 hCoV-19/England/CAMB-1B89CF/2020|EPI_ISL_651673|20    ····················  ····················  ·····················  ····················  ····················  ····················
S    8540 hCoV-19/England/CAMB-1B8A08/2020|EPI_ISL_651674|20    ····················  ····················  ·····················  ····················  ····················  ····················
S    8541 hCoV-19/England/CAMB-1B8A35/2020|EPI_ISL_651675|20    ····················  ····················  ·····················  ····················  ····················  ····················
S    8542 hCoV-19/England/CAMB-1B8A80/2020|EPI_ISL_651676|20    ····················  ····················  ·····················  ····················  ····················  ····················
S    8543 hCoV-19/England/CAMB-1B8AF9/2020|EPI_ISL_651677|20    ····················  ····················  ·····················  ····················  ····················  ····················
S    8544 hCoV-19/England/CAMB-1B8B05/2020|EPI_ISL_651678|20    ····················  ····················  ·····················  ····················  ····················  ····················
S    8545 hCoV-19/England/CAMB-1B8B9C/2020|EPI_ISL_651679|20    ····················  ····················  ·····················  ····················  ····················  ····················
S    8546 hCoV-19/England/CAMB-1B8BBA/2020|EPI_ISL_651680|20    ····················  ····················  ·····················  ····················  ····················  ····················
S    8547 hCoV-19/England/CAMB-1B8C20/2020|EPI_ISL_651681|20    ····················  ····················  ·····················  ····················  ····················  ····················
S    8549 hCoV-19/England/CAMB-1B8CC6/2020|EPI_ISL_651683|20    ····················  ····················  ·····················  ····················  ····················  ····················
S    8550 hCoV-19/England/CAMB-1B8D3C/2020|EPI_ISL_651684|20    ····················  ····················  ·····················  ····················  ····················  ····················
S    8551 hCoV-19/England/CAMB-1B8D4B/2020|EPI_ISL_651685|20    ····················  ····················  ·····················  ····················  ····················  ····················
S    8552 hCoV-19/England/CAMB-1B8D87/2020|EPI_ISL_651686|20    ····················  ····················  ·····················  ····················  ····················  ····················
S    8553 hCoV-19/England/CAMB-1B8DB4/2020|EPI_ISL_651687|20    ····················  ····················  ·····················  ····················  ····················  ····················
S    8554 hCoV-19/England/CAMB-1B8DD2/2020|EPI_ISL_651688|20    ····················  ····················  ·····················  ····················  ····················  ····················
S    8555 hCoV-19/England/CAMB-1B8DF0/2020|EPI_ISL_651689|20    ····················  ····················  ·····················  ····················  ····················  ····················
S    8556 hCoV-19/England/CAMB-1B8E2A/2020|EPI_ISL_651690|20    ····················  ····················  ·····················  ····················  ····················  ····················
S    8557 hCoV-19/England/CAMB-1B8F27/2020|EPI_ISL_651691|20    ····················  ····················  ·····················  ····················  ······T·············  ····················
S    8558 hCoV-19/England/CAMB-1B8F54/2020|EPI_ISL_651692|20    ····················  ····················  ·····················  ····················  ····················  ····················
S    8559 hCoV-19/England/CAMB-1B8F63/2020|EPI_ISL_651693|20    ····················  ····················  ·····················  ····················  ····················  ····················
S    8560 hCoV-19/England/CAMB-1B8F72/2020|EPI_ISL_651694|20    ····················  ····················  ·····················  ····················  ····················  ····················
S    8561 hCoV-19/England/CAMB-1B8F81/2020|EPI_ISL_651695|20    ····················  ····················  ·····················  ····················  ····················  ····················
S    8562 hCoV-19/England/CAMB-1B9296/2020|EPI_ISL_651696|20    ····················  ····················  ·····················  ····················  ····················  ····················
S    8563 hCoV-19/England/CAMB-1B92C3/2020|EPI_ISL_651697|20    ····················  ····················  ·····················  ····················  ······T·············  ····················
S    8564 hCoV-19/England/CAMB-1B92D2/2020|EPI_ISL_651698|20    ····················  ····················  ·····················  ····················  ······T·············  ····················
S    8565 hCoV-19/England/CAMB-1B930C/2020|EPI_ISL_651699|20    ····················  ····················  ·····················  ····················  ······T·············  ····················
S    8566 hCoV-19/England/QEUH-AAE3C5/2020|EPI_ISL_652973|20    ····················  ····················  ·····················  ····················  ····················  ····················
S    8567 hCoV-19/England/QEUH-AAE5DE/2020|EPI_ISL_652974|20    ····················  ····················  ·····················  ····················  ····················  ····················
S    8568 hCoV-19/England/QEUH-AAE732/2020|EPI_ISL_652975|20    ····················  ····················  ·····················  ····················  ····················  ····················
S    8569 hCoV-19/England/QEUH-AAE741/2020|EPI_ISL_652976|20    ····················  ····················  ·····················  ····················  ····················  ····················
S    8570 hCoV-19/England/QEUH-AAEA0C/2020|EPI_ISL_652977|20    ····················  ····················  ·····················  ····················  ····················  ····················
S    8571 hCoV-19/England/QEUH-AAEA2A/2020|EPI_ISL_652978|20    ····················  ····················  ·····················  ····················  ····················  ····················
S    8572 hCoV-19/England/QEUH-AAEA39/2020|EPI_ISL_652979|20    ····················  ····················  ·····················  ····················  ····················  ····················
S    8573 hCoV-19/England/QEUH-AAEA75/2020|EPI_ISL_652980|20    ····················  ····················  ·····················  ····················  ····················  ····················
S    8574 hCoV-19/England/QEUH-AAEAA2/2020|EPI_ISL_652981|20    ····················  ····················  ·····················  ····················  ····················  ····················
S    8575 hCoV-19/England/QEUH-AAEADF/2020|EPI_ISL_652982|20    ····················  ····················  ·····················  ····················  ····················  ····················
S    8576 hCoV-19/England/QEUH-AAEAFD/2020|EPI_ISL_652983|20    ····················  ····················  ·····················  ····················  ····················  ····················
S    8577 hCoV-19/England/QEUH-AAEB09/2020|EPI_ISL_652984|20    ····················  ····················  ·····················  ····················  ····················  ····················
S    8578 hCoV-19/England/QEUH-AAEB18/2020|EPI_ISL_652985|20    ····················  ····················  ·····················  ····················  ····················  ····················
S    8579 hCoV-19/England/QEUH-AAEB36/2020|EPI_ISL_652986|20    ····················  ····················  ·····················  ····················  ····················  ····················
S    8580 hCoV-19/England/QEUH-AAEB72/2020|EPI_ISL_652987|20    ····················  ····················  ·····················  ····················  ····················  ····················
S    8581 hCoV-19/England/QEUH-AAEBAF/2020|EPI_ISL_652988|20    ····················  ····················  ·····················  ····················  ····················  ····················
S    8582 hCoV-19/England/QEUH-AAEBBE/2020|EPI_ISL_652989|20    ····················  ····················  ·····················  ····················  ····················  ····················
S    8583 hCoV-19/England/QEUH-AAEBCD/2020|EPI_ISL_652990|20    ····················  ····················  ·····················  ····················  ····················  ····················
S    8584 hCoV-19/England/QEUH-AAEBDC/2020|EPI_ISL_652991|20    ····················  ····················  ·····················  ····················  ····················  ····················
S    8585 hCoV-19/England/QEUH-AAEC51/2020|EPI_ISL_652992|20    ····················  ····················  ·····················  ····················  ····················  ····················
S    8586 hCoV-19/England/QEUH-AAEC60/2020|EPI_ISL_652993|20    ····················  ····················  ·····················  ····················  ····················  ····················
S    8587 hCoV-19/England/QEUH-AAEC7F/2020|EPI_ISL_652994|20    ····················  ····················  ·····················  ····················  ····················  ····················
S    8588 hCoV-19/England/QEUH-AAEC8E/2020|EPI_ISL_652995|20    ····················  ····················  ·····················  ····················  ····················  ····················
S    8589 hCoV-19/England/QEUH-AAEC9D/2020|EPI_ISL_652996|20    ····················  ····················  ·····················  ····················  ····················  ····················
S    8590 hCoV-19/England/QEUH-AAECAC/2020|EPI_ISL_652997|20    ····················  ····················  ·····················  ····················  ····················  ····················
S    8591 hCoV-19/England/QEUH-AAECBB/2020|EPI_ISL_652998|20    ····················  ····················  ·····················  ····················  ······T·············  ····················
S    8592 hCoV-19/England/QEUH-AAECF7/2020|EPI_ISL_652999|20    ····················  ····················  ·····················  ····················  ····················  ····················
S    8593 hCoV-19/England/QEUH-B6C16E/2020|EPI_ISL_655150|20    ····················  ····················  ·····················  ····················  ····················  ····················
S    8594 hCoV-19/England/QEUH-B69ED3/2020|EPI_ISL_655151|20    ····················  ····················  ·····················  ····················  ····················  ····················
S    8595 hCoV-19/England/QEUH-B69BBE/2020|EPI_ISL_655152|20    ····················  ····················  ·····················  ····················  ····················  ····················
S    8596 hCoV-19/England/QEUH-B681CC/2020|EPI_ISL_655153|20    ····················  ····················  ·····················  ····················  ····················  ····················
```

| | | | | | | | |
|---|---|---|---|---|---|---|---|
| S | 8597 hCoV-19/England/QEUH-B6BEB3/2020\|EPI_ISL_655154\|20 | ···················· | ···················· | ····················· | ···················· | ···················· | ···················· |
| S | 8598 hCoV-19/England/QEUH-B69BDC/2020\|EPI_ISL_655155\|20 | ···················· | ·A·················· | ····················· | ···················· | ···················· | ···················· |
| S | 8599 hCoV-19/England/QEUH-B6877F/2020\|EPI_ISL_655156\|20 | ···················· | ···················· | ····················· | ···················· | ···················· | ···················· |
| S | 8600 hCoV-19/England/QEUH-B6C8C3/2020\|EPI_ISL_655157\|20 | ···················· | ···················· | ····················· | ···················· | ···T················ | ···················· |
| S | 8601 hCoV-19/England/QEUH-B6C9A2/2020\|EPI_ISL_655158\|20 | ···················· | ···················· | ····················· | ···················· | ···················· | ··A················· |
| S | 8602 hCoV-19/England/QEUH-B6AC50/2020\|EPI_ISL_655160\|20 | ···················· | ···················· | ····················· | ···················· | ···················· | ···················· |
| S | 8603 hCoV-19/England/QEUH-B69F94/2020\|EPI_ISL_655162\|20 | ···················· | ···················· | ····················· | ···················· | ···················· | ···················· |
| S | 8604 hCoV-19/England/QEUH-B6C8D2/2020\|EPI_ISL_655163\|20 | ···················· | ···················· | ····················· | ···················· | ···················· | ···················· |
| S | 8605 hCoV-19/England/QEUH-B6BE77/2020\|EPI_ISL_655164\|20 | ···················· | ···················· | ····················· | ···················· | ···················· | ···················· |
| S | 8606 hCoV-19/England/QEUH-B6C17D/2020\|EPI_ISL_655165\|20 | ···················· | ···················· | ····················· | ···················· | ···················· | ···················· |
| S | 8607 hCoV-19/England/QEUH-B6ACAB/2020\|EPI_ISL_655166\|20 | ···················· | ···················· | ····················· | ···················· | ···················· | ···················· |
| S | 8608 hCoV-19/England/QEUH-B68AB2/2020\|EPI_ISL_655169\|20 | ···················· | ···················· | ····················· | ···················· | ···················· | ···················· |
| S | 8609 hCoV-19/England/ALDP-B63A6C/2020\|EPI_ISL_656400\|20 | ···················· | ···················· | ····················· | ···················· | ···················· | ···················· |
| S | 8610 hCoV-19/England/ALDP-B6348C/2020\|EPI_ISL_656401\|20 | ···················· | ···················· | ····················· | ···················· | ···················· | ···················· |
| S | 8611 hCoV-19/England/ALDP-B635F2/2020\|EPI_ISL_656402\|20 | ···················· | ···················· | ····················· | ···················· | ···················· | ···················· |
| S | 8612 hCoV-19/England/ALDP-B62739/2020\|EPI_ISL_656403\|20 | ···················· | ·A·················· | ····················· | ···················· | ···················· | ··A················· |
| S | 8613 hCoV-19/England/ALDP-B63404/2020\|EPI_ISL_656404\|20 | ···················· | ···················· | ····················· | ···················· | ···················· | ···················· |
| S | 8614 hCoV-19/England/ALDP-B622FC/2020\|EPI_ISL_656405\|20 | ···················· | ···················· | ····················· | ···················· | ···················· | ···················· |
| S | 8615 hCoV-19/England/ALDP-B629F7/2020\|EPI_ISL_656406\|20 | ···················· | ···················· | ····················· | ···················· | ···················· | ···················· |
| S | 8616 hCoV-19/England/ALDP-B62098/2020\|EPI_ISL_656407\|20 | ···················· | ···················· | ····················· | ···················· | ···················· | ···················· |
| S | 8617 hCoV-19/England/ALDP-B632FB/2020\|EPI_ISL_656408\|20 | ···················· | ···················· | ····················· | ···················· | ···················· | ···················· |
| S | 8618 hCoV-19/England/ALDP-B639F6/2020\|EPI_ISL_656410\|20 | ···················· | ···················· | ····················· | ···················· | ···················· | ···················· |
| S | 8619 hCoV-19/England/ALDP-B63659/2020\|EPI_ISL_656411\|20 | ···················· | ···················· | ····················· | ···················· | ···················· | ··A················· |
| S | 8620 hCoV-19/England/ALDP-B627C0/2020\|EPI_ISL_656412\|20 | ···················· | ···················· | ····················· | ···················· | ···················· | ···················· |
| S | 8621 hCoV-19/England/ALDP-B63C39/2020\|EPI_ISL_656413\|20 | ···················· | ···················· | ····················· | ···················· | ·····T·············· | ···················· |
| S | 8622 hCoV-19/England/ALDP-B62933/2020\|EPI_ISL_656414\|20 | ···················· | ···················· | ····················· | ···················· | ···················· | ···················· |
| S | 8623 hCoV-19/England/ALDP-B634F5/2020\|EPI_ISL_656480\|20 | ···················· | ···················· | ····················· | ···················· | ···················· | ···················· |
| S | 8624 hCoV-19/England/ALDP-B62915/2020\|EPI_ISL_656481\|20 | ···················· | ···················· | ····················· | ···················· | ···················· | ···················· |
| S | 8625 hCoV-19/England/ALDP-B623DB/2020\|EPI_ISL_656482\|20 | ···················· | ···················· | ····················· | ···················· | ···················· | ···················· |
| S | 8626 hCoV-19/England/ALDP-B6370B/2020\|EPI_ISL_656483\|20 | ···················· | ···················· | ····················· | ···················· | ···················· | ···················· |
| S | 8627 hCoV-19/England/ALDP-B63CC0/2020\|EPI_ISL_656484\|20 | ···················· | ···················· | ····················· | ···················· | ···················· | ···················· |
| S | 8628 hCoV-19/England/ALDP-B62836/2020\|EPI_ISL_656486\|20 | ···················· | ···················· | ····················· | ···················· | ···················· | ···················· |
| S | 8629 hCoV-19/England/ALDP-B634C8/2020\|EPI_ISL_656487\|20 | ···················· | ···················· | ····················· | ···················· | ···················· | ···················· |
| S | 8630 hCoV-19/England/ALDP-B623CC/2020\|EPI_ISL_656488\|20 | ···················· | ···················· | ····················· | ···················· | ···················· | ···················· |
| S | 8631 hCoV-19/England/ALDP-B62678/2020\|EPI_ISL_656489\|20 | ···················· | ·A·················· | ····················· | ···················· | ···················· | ··A················· |
| S | 8632 hCoV-19/England/ALDP-B62450/2020\|EPI_ISL_656490\|20 | ···················· | ···················· | ····················· | ···················· | ···················· | ···················· |
| S | 8633 hCoV-19/England/ALDP-B6299D/2020\|EPI_ISL_656491\|20 | ···················· | ···················· | ····················· | ···················· | ···················· | ···················· |
| S | 8634 hCoV-19/England/ALDP-B63923/2020\|EPI_ISL_656492\|20 | ···················· | ···················· | ····················· | ···················· | ···················· | ···················· |
| S | 8635 hCoV-19/England/ALDP-B6205C/2020\|EPI_ISL_656494\|20 | ···················· | ···················· | ····················· | ···················· | ···················· | ···················· |
| S | 8636 hCoV-19/England/ALDP-B629AC/2020\|EPI_ISL_656495\|20 | ···················· | ···················· | ····················· | ···················· | ···················· | ···················· |
| S | 8637 hCoV-19/England/ALDP-B63B5A/2020\|EPI_ISL_656497\|20 | ···················· | ···················· | ····················· | ···················· | ···················· | ···················· |
| S | 8638 hCoV-19/England/ALDP-B635E3/2020\|EPI_ISL_656499\|20 | ···················· | ···················· | ····················· | ···················· | ···················· | ···················· |
| S | 8639 hCoV-19/England/MILK-B564E6/2020\|EPI_ISL_657700\|20 | ···················· | ·········T·········· | ····················· | ···················· | ···················· | ···················· |
| S | 8640 hCoV-19/England/CAMC-B530D6/2020\|EPI_ISL_657701\|20 | ···················· | ···················· | ····················· | ···················· | ···················· | ···················· |
| S | 8641 hCoV-19/England/MILK-B55960/2020\|EPI_ISL_657703\|20 | ···················· | ···················· | ····················· | ···················· | ···················· | ···················· |
| S | 8642 hCoV-19/England/MILK-B55A03/2020\|EPI_ISL_657705\|20 | ···················· | ···················· | ····················· | ···················· | ···················· | ···················· |
| S | 8643 hCoV-19/England/MILK-B55872/2020\|EPI_ISL_657706\|20 | ···················· | ···················· | ····················· | ···················· | ···················· | ···················· |
| S | 8644 hCoV-19/England/CAMC-B531A6/2020\|EPI_ISL_657707\|20 | ···················· | ···················· | ····················· | ···················· | ···················· | ···················· |
| S | 8645 hCoV-19/England/CAMC-B52B8B/2020\|EPI_ISL_657708\|20 | ···················· | ·A·················· | ····················· | ···················· | ···················· | ··A················· |
| S | 8646 hCoV-19/England/MILK-B561B2/2020\|EPI_ISL_657709\|20 | ···················· | ···················· | ····················· | ···················· | ···················· | ···················· |
| S | 8647 hCoV-19/England/MILK-B5653E/2020\|EPI_ISL_657710\|20 | ···················· | ···················· | ····················· | ···················· | ···················· | ···················· |
| S | 8648 hCoV-19/England/MILK-B5613A/2020\|EPI_ISL_657711\|20 | ···················· | ···················· | ····················· | ···················· | ···················· | ···················· |
| S | 8649 hCoV-19/England/MILK-B5505C/2020\|EPI_ISL_657712\|20 | ···················· | ···················· | ····················· | ···················· | ···················· | ···················· |
| S | 8650 hCoV-19/England/MILK-B550C5/2020\|EPI_ISL_657714\|20 | ···················· | ···················· | ····················· | ···················· | ···················· | ···················· |
| S | 8651 hCoV-19/England/MILK-B54E71/2020\|EPI_ISL_657715\|20 | ···················· | ···················· | ····················· | ···················· | ···················· | ···················· |
| S | 8652 hCoV-19/England/MILK-B552A1/2020\|EPI_ISL_657716\|20 | ···················· | ···················· | ····················· | ···················· | ···················· | ···················· |
| S | 8653 hCoV-19/England/MILK-B54B7A/2020\|EPI_ISL_657717\|20 | ···················· | ···················· | ····················· | ···················· | ···················· | ···················· |
| S | 8654 hCoV-19/England/MILK-B54C0E/2020\|EPI_ISL_657718\|20 | ···················· | ···················· | ····················· | ···················· | ···················· | ···················· |
| S | 8655 hCoV-19/England/CAMC-B515F7/2020\|EPI_ISL_657721\|20 | ···················· | ···················· | ····················· | ···················· | ···················· | ···················· |
| S | 8656 hCoV-19/England/MILK-B54D38/2020\|EPI_ISL_657722\|20 | ···················· | ···················· | ····················· | ···················· | ···················· | ···················· |
| S | 8657 hCoV-19/England/CAMC-B520F5/2020\|EPI_ISL_657723\|20 | ···················· | ···················· | ····················· | ···················· | ···················· | ···················· |
| S | 8658 hCoV-19/England/CAMC-B52040/2020\|EPI_ISL_657724\|20 | ···················· | ···················· | ····················· | ···················· | ···················· | ···················· |
| S | 8659 hCoV-19/England/CAMC-B51E65/2020\|EPI_ISL_657725\|20 | ···················· | ···················· | ····················· | ···················· | ···················· | ···················· |
| S | 8660 hCoV-19/England/CAMC-B5164F/2020\|EPI_ISL_657726\|20 | ···················· | ·A·················· | ····················· | ···················· | ···················· | ··A················· |
| S | 8661 hCoV-19/England/MILK-B55599/2020\|EPI_ISL_657727\|20 | ···················· | ···················· | ····················· | ···················· | ···················· | ···················· |
| S | 8662 hCoV-19/England/CAMC-B51FCC/2020\|EPI_ISL_657728\|20 | ···················· | ·A·················· | ····················· | ···················· | ···················· | ··A················· |
| S | 8663 hCoV-19/England/MILK-B54B98/2020\|EPI_ISL_657729\|20 | ···················· | ···················· | ····················· | ···················· | ···················· | ···················· |
| S | 8664 hCoV-19/England/MILK-B54D0B/2020\|EPI_ISL_657730\|20 | ···················· | ···················· | ····················· | ···················· | ···················· | ···················· |
| S | 8665 hCoV-19/England/MILK-B554F6/2020\|EPI_ISL_657732\|20 | ···················· | ···················· | ····················· | ···················· | ···················· | ···················· |
| S | 8666 hCoV-19/England/MILK-B55423/2020\|EPI_ISL_657733\|20 | ···················· | ···················· | ····················· | ···················· | ···················· | ···················· |
| S | 8667 hCoV-19/England/MILK-B5548D/2020\|EPI_ISL_657734\|20 | ···················· | ···················· | ····················· | ···················· | ···················· | ···················· |
| S | 8668 hCoV-19/England/MILK-B54EF9/2020\|EPI_ISL_657735\|20 | ···················· | ···················· | ····················· | ···················· | ···················· | ···················· |
| S | 8669 hCoV-19/England/MILK-B5554E/2020\|EPI_ISL_657738\|20 | ···················· | ···················· | ····················· | ···················· | ···················· | ···················· |
| S | 8670 hCoV-19/England/CAMC-B51490/2020\|EPI_ISL_657739\|20 | ···················· | ···················· | ····················· | ···················· | ···················· | ···················· |
| S | 8671 hCoV-19/England/MILK-B54EAE/2020\|EPI_ISL_657740\|20 | ···················· | ···················· | ····················· | ···················· | ···················· | ···················· |
| S | 8672 hCoV-19/England/MILK-B54E80/2020\|EPI_ISL_657741\|20 | ···················· | ···················· | ····················· | ···················· | ···················· | ···················· |
| S | 8673 hCoV-19/England/MILK-B555A8/2020\|EPI_ISL_657742\|20 | ···················· | ···················· | ····················· | ···················· | ···················· | ···················· |
| S | 8674 hCoV-19/England/MILK-B54CFF/2020\|EPI_ISL_657743\|20 | ···················· | ···················· | ····················· | ···················· | ···················· | ···················· |
| S | 8675 hCoV-19/England/CAMC-B5115D/2020\|EPI_ISL_657744\|20 | ···················· | ···················· | ····················· | ···················· | ···················· | ···················· |
| S | 8676 hCoV-19/England/MILK-B55159/2020\|EPI_ISL_657745\|20 | ···················· | ···················· | ····················· | ···················· | ···················· | ···················· |

| | | | | | | | |
|---|---|---|---|---|---|---|---|
| S | 8677 hCoV-19/England/CAMC-B51296/2020\|EPI_ISL_657746\|20 | ···················· | ···················· | ···················· | ···················· | ···················· | ···················· |
| S | 8678 hCoV-19/England/CAMC-B52013/2020\|EPI_ISL_657790\|20 | ···················· | ···················· | ···················· | ···················· | ···················· | ···················· |
| S | 8679 hCoV-19/USA/WI-GMF-28010/2020\|EPI_ISL_661000\|2020- | ···················· | ···················· | ···················· | ···················· | ···················· | ···················· |
| S | 8680 hCoV-19/USA/WI-GMF-28025/2020\|EPI_ISL_661001\|2020- | ···················· | ···················· | ···················· | ···················· | ···················· | ···················· |
| S | 8681 hCoV-19/USA/WI-GMF-28057/2020\|EPI_ISL_661002\|2020- | ···················· | ···················· | ···················· | ···················· | ···················· | ···················· |
| S | 8682 hCoV-19/USA/WI-GMF-28096/2020\|EPI_ISL_661003\|2020- | ···················· | ···················· | ···················· | ···················· | ···················· | ···················· |
| S | 8683 hCoV-19/USA/WI-GMF-28099/2020\|EPI_ISL_661004\|2020- | ···················· | ···················· | ···················· | ···················· | ···················· | ···················· |
| S | 8684 hCoV-19/USA/WI-GMF-28108/2020\|EPI_ISL_661005\|2020- | ···················· | ···················· | ···················· | ···················· | ···················· | ···················· |
| S | 8685 hCoV-19/USA/MN-GMF-28113/2020\|EPI_ISL_661006\|2020- | ···················· | ···················· | ···················· | ···················· | ···················· | ···················· |
| S | 8686 hCoV-19/USA/WI-GMF-28119/2020\|EPI_ISL_661007\|2020- | ···················· | ···················· | ···················· | ···················· | ···················· | ···················· |
| S | 8687 hCoV-19/USA/WI-GMF-28137/2020\|EPI_ISL_661008\|2020- | ···················· | ···················· | ···················· | ···················· | ···················· | ···················· |
| S | 8688 hCoV-19/USA/WI-GMF-28150/2020\|EPI_ISL_661009\|2020- | ···················· | ···················· | ···················· | ···················· | ···················· | ···················· |
| S | 8689 hCoV-19/USA/WI-GMF-28179/2020\|EPI_ISL_661010\|2020- | ···················· | ···················· | ···················· | ···················· | ···················· | ···················· |
| S | 8690 hCoV-19/USA/WI-GMF-28185/2020\|EPI_ISL_661011\|2020- | ···················· | ···················· | ···················· | ···················· | ···················· | ···················· |
| S | 8691 hCoV-19/USA/WI-GMF-28220/2020\|EPI_ISL_661012\|2020- | ···················· | ···················· | ···················· | ···················· | ···················· | ···················· |
| S | 8692 hCoV-19/USA/MN-GMF-28226/2020\|EPI_ISL_661013\|2020- | ···················· | ···················· | ···················· | ···················· | ···················· | ···················· |
| S | 8693 hCoV-19/USA/WI-GMF-28325/2020\|EPI_ISL_661014\|2020- | ···················· | ···················· | ···················· | ···················· | ···················· | ···················· |
| S | 8694 hCoV-19/USA/WI-GMF-28356/2020\|EPI_ISL_661015\|2020- | ···················· | ···················· | ···················· | ···················· | ···················· | ···················· |
| S | 8695 hCoV-19/USA/WI-GMF-28395/2020\|EPI_ISL_661016\|2020- | ···················· | ···················· | ···················· | ···················· | ···················· | ···················· |
| S | 8696 hCoV-19/USA/WI-GMF-28399/2020\|EPI_ISL_661017\|2020- | ···················· | ···················· | ···················· | ···················· | ···················· | ···················· |
| S | 8697 hCoV-19/USA/IA-GMF-28410/2020\|EPI_ISL_661018\|2020- | ···················· | ···················· | ···················· | ···················· | ···················· | ···················· |
| S | 8698 hCoV-19/USA/WI-GMF-28414/2020\|EPI_ISL_661019\|2020- | ···················· | ···················· | ···················· | ···················· | ···················· | ···················· |
| S | 8699 hCoV-19/USA/WI-GMF-28421/2020\|EPI_ISL_661020\|2020- | ···················· | ···················· | ···················· | ···················· | ···················· | ···················· |
| S | 8700 hCoV-19/USA/IA-GMF-28426/2020\|EPI_ISL_661021\|2020- | ···················· | ···················· | ···················· | ···················· | ···················· | ···················· |
| S | 8701 hCoV-19/USA/WI-GMF-30007/2020\|EPI_ISL_661022\|2020- | ···················· | ···················· | ···················· | ···················· | ···················· | ···················· |
| S | 8702 hCoV-19/USA/WI-GMF-30040/2020\|EPI_ISL_661023\|2020- | ···················· | ···················· | ···················· | ···················· | ···················· | ···················· |
| S | 8703 hCoV-19/USA/WI-GMF-30052/2020\|EPI_ISL_661024\|2020- | ···················· | ···················· | ···················· | ···················· | ···················· | ···················· |
| S | 8704 hCoV-19/USA/WI-GMF-30474/2020\|EPI_ISL_661035\|2020- | ···················· | ···················· | ···················· | ···················· | ···················· | ···················· |
| S | 8705 hCoV-19/England/QEUH-B682C9/2020\|EPI_ISL_655101\|20 | ···················· | ·A·················· | ···················· | ···················· | ···················· | ··A················· |
| S | 8706 hCoV-19/England/QEUH-B685A2/2020\|EPI_ISL_655102\|20 | ···················· | ···················· | ···················· | ···················· | ···················· | ···················· |
| S | 8707 hCoV-19/England/QEUH-B69732/2020\|EPI_ISL_655103\|20 | ···················· | ···················· | ···················· | ···················· | ···················· | ···················· |
| S | 8708 hCoV-19/England/QEUH-B6C984/2020\|EPI_ISL_655104\|20 | ···················· | ···················· | ···················· | ···················· | ·····T·············· | ···················· |
| S | 8709 hCoV-19/England/QEUH-B68B28/2020\|EPI_ISL_655106\|20 | ···················· | ···················· | ···················· | ···················· | ···················· | ···················· |
| S | 8710 hCoV-19/England/QEUH-B69F49/2020\|EPI_ISL_655109\|20 | ···················· | ···················· | ···················· | ···················· | ···················· | ···················· |
| S | 8711 hCoV-19/England/QEUH-B65E04/2020\|EPI_ISL_655110\|20 | ···················· | ···················· | ···················· | ···················· | ···················· | ···················· |
| S | 8712 hCoV-19/England/QEUH-B6C0CB/2020\|EPI_ISL_655112\|20 | ···················· | ···················· | ···················· | ···················· | ···················· | ···················· |
| S | 8713 hCoV-19/England/QEUH-B6C122/2020\|EPI_ISL_655113\|20 | ···················· | ···················· | ···················· | ···················· | ···················· | ···················· |
| S | 8714 hCoV-19/England/QEUH-B65D61/2020\|EPI_ISL_655114\|20 | ···················· | ···················· | ···················· | ···················· | ···················· | ···················· |
| S | 8715 hCoV-19/England/QEUH-B67D23/2020\|EPI_ISL_655116\|20 | ···················· | ···················· | ···················· | ···················· | ···················· | ···················· |
| S | 8716 hCoV-19/England/QEUH-B6AC23/2020\|EPI_ISL_655117\|20 | ···················· | ·A·················· | ···················· | ···················· | ···················· | ···················· |
| S | 8717 hCoV-19/England/QEUH-B68AC1/2020\|EPI_ISL_655118\|20 | ···················· | ···················· | ···················· | ···················· | ···················· | ···················· |
| S | 8718 hCoV-19/England/QEUH-B682E7/2020\|EPI_ISL_655119\|20 | ···················· | ···················· | ···················· | ···················· | ···················· | ···················· |
| S | 8719 hCoV-19/England/QEUH-B6BE2C/2020\|EPI_ISL_655120\|20 | ···················· | ···················· | ···················· | ···················· | ···················· | ···················· |
| S | 8720 hCoV-19/England/QEUH-B68B64/2020\|EPI_ISL_655121\|20 | ···················· | ···················· | ···················· | ···················· | ···················· | ···················· |
| S | 8721 hCoV-19/England/QEUH-B65DCB/2020\|EPI_ISL_655122\|20 | ···················· | ···················· | ···················· | ···················· | ···················· | ···················· |
| S | 8722 hCoV-19/England/QEUH-B6AC7E/2020\|EPI_ISL_655123\|20 | ···················· | ···················· | ···················· | ···················· | ···················· | ···················· |
| S | 8723 hCoV-19/England/QEUH-B6AC6F/2020\|EPI_ISL_655125\|20 | ···················· | ···················· | ···················· | ···················· | ···················· | ···················· |
| S | 8724 hCoV-19/England/QEUH-B6A27C/2020\|EPI_ISL_655126\|20 | ···················· | ···················· | ···················· | ···················· | ···················· | ···················· |
| S | 8725 hCoV-19/England/QEUH-B69BCD/2020\|EPI_ISL_655127\|20 | ···················· | ···················· | ···················· | ···················· | ···················· | ···················· |
| S | 8726 hCoV-19/England/QEUH-B6AC14/2020\|EPI_ISL_655129\|20 | ···················· | ···················· | ···················· | ···················· | ···················· | ···················· |
| S | 8727 hCoV-19/England/QEUH-B67D50/2020\|EPI_ISL_655130\|20 | ···················· | ···················· | ···················· | ···················· | ···················· | ···················· |
| S | 8728 hCoV-19/England/QEUH-B66A4B/2020\|EPI_ISL_655132\|20 | ···················· | ···················· | ···················· | ···················· | ···················· | ···················· |
| S | 8729 hCoV-19/England/QEUH-B67DAB/2020\|EPI_ISL_655133\|20 | ···················· | ···················· | ···················· | ···················· | ···················· | ···················· |
| S | 8730 hCoV-19/England/QEUH-B68A94/2020\|EPI_ISL_655134\|20 | ···T················ | ···················· | ···················· | ···················· | ···················· | ···················· |
| S | 8731 hCoV-19/England/QEUH-B6C070/2020\|EPI_ISL_655135\|20 | ···················· | ···················· | ···················· | ···················· | ···················· | ···················· |
| S | 8732 hCoV-19/England/QEUH-B69C7F/2020\|EPI_ISL_655136\|20 | ···················· | ···················· | ···················· | ···················· | ···················· | ···················· |
| S | 8733 hCoV-19/England/QEUH-B684F0/2020\|EPI_ISL_655137\|20 | ···················· | ···················· | ···················· | ···················· | ···················· | ···················· |
| S | 8734 hCoV-19/England/QEUH-B6C429/2020\|EPI_ISL_655138\|20 | ···················· | ···················· | ···················· | ···················· | ···················· | ···················· |
| S | 8735 hCoV-19/England/QEUH-B65D8F/2020\|EPI_ISL_655139\|20 | ···················· | ···················· | ···················· | ···················· | ···················· | ···················· |
| S | 8736 hCoV-19/England/QEUH-B65D34/2020\|EPI_ISL_655140\|20 | ···················· | ···················· | ···················· | ···················· | ···················· | ···················· |
| S | 8737 hCoV-19/England/QEUH-B6969F/2020\|EPI_ISL_655142\|20 | ···················· | ···················· | ···················· | ···················· | ···················· | ···················· |
| S | 8738 hCoV-19/England/QEUH-B69671/2020\|EPI_ISL_655143\|20 | ···················· | ···················· | ···················· | ···················· | ···················· | ···················· |
| S | 8739 hCoV-19/England/QEUH-B6C438/2020\|EPI_ISL_655144\|20 | ···················· | ···················· | ···················· | ···················· | ···················· | ···················· |
| S | 8740 hCoV-19/England/QEUH-B65CEC/2020\|EPI_ISL_655145\|20 | ···················· | ···················· | ···················· | ···················· | ···················· | ···················· |
| S | 8741 hCoV-19/England/QEUH-B6BA55/2020\|EPI_ISL_655146\|20 | ···················· | ···················· | ···················· | ···················· | ···················· | ···················· |
| S | 8742 hCoV-19/England/QEUH-B6A26D/2020\|EPI_ISL_655147\|20 | ···················· | ·A·················· | ···················· | ···················· | ···················· | ··A················· |
| S | 8743 hCoV-19/England/QEUH-B67DD8/2020\|EPI_ISL_655148\|20 | ···················· | ···················· | ···················· | ···················· | ···················· | ···················· |
| S | 8744 hCoV-19/England/QEUH-B6B9D0/2020\|EPI_ISL_655149\|20 | ···················· | ···················· | ···················· | ···················· | ···················· | ···················· |
| S | 8745 hCoV-19/England/ALDP-B63246/2020\|EPI_ISL_656416\|20 | ···················· | ···················· | ···················· | ···················· | ···················· | ···················· |
| S | 8746 hCoV-19/England/ALDP-B63C75/2020\|EPI_ISL_656417\|20 | ···················· | ···················· | ···················· | ···················· | ···················· | ···················· |
| S | 8747 hCoV-19/England/ALDP-B62748/2020\|EPI_ISL_656418\|20 | ···················· | ···················· | ···················· | ···················· | ···················· | ··A················· |
| S | 8748 hCoV-19/England/ALDP-B63325/2020\|EPI_ISL_656419\|20 | ···················· | ···················· | ···················· | ···················· | ···················· | ···················· |
| S | 8749 hCoV-19/England/ALDP-B62854/2020\|EPI_ISL_656420\|20 | ···················· | ···················· | ···················· | ···················· | ···················· | ···················· |
| S | 8750 hCoV-19/England/ALDP-B63C2A/2020\|EPI_ISL_656422\|20 | ···················· | ···················· | ···················· | ···················· | ···················· | ···················· |
| S | 8751 hCoV-19/England/ALDP-B622CF/2020\|EPI_ISL_656423\|20 | ···················· | ···················· | ···················· | ···················· | ···················· | ···················· |
| S | 8752 hCoV-19/England/ALDP-B6272A/2020\|EPI_ISL_656424\|20 | ···················· | ···················· | ···················· | ···················· | ···················· | ···················· |
| S | 8753 hCoV-19/England/ALDP-B63440/2020\|EPI_ISL_656425\|20 | ···················· | ···················· | ···················· | ···················· | ···················· | ···················· |
| S | 8754 hCoV-19/England/ALDP-B638CC/2020\|EPI_ISL_656426\|20 | ···················· | ···················· | ···················· | ···················· | ···················· | ···················· |
| S | 8755 hCoV-19/England/ALDP-B62B1F/2020\|EPI_ISL_656427\|20 | ···················· | ···················· | ···················· | ···················· | ···················· | ···················· |
| S | 8756 hCoV-19/England/ALDP-B63668/2020\|EPI_ISL_656430\|20 | ···················· | ·A·················· | ···················· | ···················· | ···················· | ···················· |

| | | | | | | | |
|---|---|---|---|---|---|---|---|
| S | 8757 hCoV-19/England/ALDP-B63370/2020\|EPI_ISL_656431\|20 | ···················· | ·A·················· | ····················· | ···················· | ···················· | ···A················ |
| S | 8758 hCoV-19/England/ALDP-B62335/2020\|EPI_ISL_656432\|20 | ···················· | ···················· | ····················· | ···················· | ···················· | ···················· |
| S | 8759 hCoV-19/England/ALDP-B62845/2020\|EPI_ISL_656433\|20 | ···················· | ···················· | ····················· | ···················· | ···················· | ···················· |
| S | 8760 hCoV-19/England/ALDP-B62010/2020\|EPI_ISL_656434\|20 | ···················· | ···················· | ····················· | ···················· | ···················· | ···················· |
| S | 8761 hCoV-19/England/ALDP-B62414/2020\|EPI_ISL_656435\|20 | ···················· | ···················· | ····················· | ···················· | ···················· | ···················· |
| S | 8762 hCoV-19/England/ALDP-B626E1/2020\|EPI_ISL_656436\|20 | ···················· | ···················· | ····················· | ···················· | ···················· | ···················· |
| S | 8763 hCoV-19/England/ALDP-B63B4B/2020\|EPI_ISL_656437\|20 | ···················· | ···················· | ····················· | ···················· | ···················· | ···················· |
| S | 8764 hCoV-19/England/ALDP-B63A20/2020\|EPI_ISL_656439\|20 | ···················· | ···················· | ····················· | ···················· | ···················· | ···················· |
| S | 8765 hCoV-19/England/ALDP-B62818/2020\|EPI_ISL_656440\|20 | ···················· | ···················· | ····················· | ···················· | ···················· | ···················· |
| S | 8766 hCoV-19/England/ALDP-B62423/2020\|EPI_ISL_656442\|20 | ···················· | ···················· | ····················· | ···················· | ···················· | ···················· |
| S | 8767 hCoV-19/England/ALDP-B6213B/2020\|EPI_ISL_656443\|20 | ···················· | ···················· | ····················· | ···················· | ······T············· | ···················· |
| S | 8768 hCoV-19/England/ALDP-B632A0/2020\|EPI_ISL_656444\|20 | ···················· | ···················· | ····················· | ···················· | ···················· | ···················· |
| S | 8769 hCoV-19/England/ALDP-B62AA9/2020\|EPI_ISL_656445\|20 | ···················· | ···················· | ····················· | ···················· | ···················· | ···················· |
| S | 8770 hCoV-19/England/ALDP-B635D4/2020\|EPI_ISL_656446\|20 | ···················· | ···················· | ····················· | ···················· | ···················· | ···················· |
| S | 8771 hCoV-19/England/ALDP-B63808/2020\|EPI_ISL_656447\|20 | ···················· | ···················· | ····················· | ···················· | ···················· | ···················· |
| S | 8772 hCoV-19/England/ALDP-B63CA2/2020\|EPI_ISL_656448\|20 | ···················· | ···················· | ····················· | ···················· | ···················· | ···················· |
| S | 8773 hCoV-19/England/ALDP-B63C93/2020\|EPI_ISL_656449\|20 | ···················· | ···················· | ····················· | ···················· | ···················· | ···················· |
| S | 8774 hCoV-19/England/ALDP-B63CFD/2020\|EPI_ISL_656450\|20 | ·······C············ | ···················· | ····················· | ···················· | ···················· | ···················· |
| S | 8775 hCoV-19/England/ALDP-B624BA/2020\|EPI_ISL_656451\|20 | ···················· | ···················· | ····················· | ···················· | ···················· | ···················· |
| S | 8776 hCoV-19/England/ALDP-B63853/2020\|EPI_ISL_656452\|20 | ···················· | ···················· | ····················· | ···················· | ···················· | ···················· |
| S | 8777 hCoV-19/England/ALDP-B63282/2020\|EPI_ISL_656455\|20 | ···················· | ···················· | ····················· | ···················· | ···················· | ···················· |
| S | 8778 hCoV-19/England/ALDP-B63AF3/2020\|EPI_ISL_656456\|20 | ···················· | ···················· | ····················· | ···················· | ···················· | ···················· |
| S | 8779 hCoV-19/England/ALDP-B63932/2020\|EPI_ISL_656458\|20 | ···················· | ···················· | ····················· | ···················· | ···················· | ···················· |
| S | 8780 hCoV-19/England/ALDP-B631EF/2020\|EPI_ISL_656460\|20 | ···················· | ···················· | ····················· | ···················· | ···················· | ···················· |
| S | 8781 hCoV-19/England/ALDP-B6397E/2020\|EPI_ISL_656462\|20 | ···················· | ···················· | ····················· | ···················· | ······T············· | ···················· |
| S | 8782 hCoV-19/England/ALDP-B63A7B/2020\|EPI_ISL_656463\|20 | ···················· | ···················· | ····················· | ···················· | ···················· | ···················· |
| S | 8783 hCoV-19/England/ALDP-B620B6/2020\|EPI_ISL_656464\|20 | ···················· | ···················· | ····················· | ···················· | ···················· | ···················· |
| S | 8784 hCoV-19/England/ALDP-B637FC/2020\|EPI_ISL_656465\|20 | ···················· | ···················· | ····················· | ···················· | ···················· | ···················· |
| S | 8785 hCoV-19/England/ALDP-B628CD/2020\|EPI_ISL_656466\|20 | ···················· | ···················· | ····················· | ···················· | ···················· | ···················· |
| S | 8786 hCoV-19/England/ALDP-B6261E/2020\|EPI_ISL_656467\|20 | ···················· | ···················· | ····················· | ···················· | ···················· | ···················· |
| S | 8787 hCoV-19/England/ALDP-B628BE/2020\|EPI_ISL_656469\|20 | ···················· | ···················· | ····················· | ···················· | ···················· | ···················· |
| S | 8789 hCoV-19/England/ALDP-B633BC/2020\|EPI_ISL_656472\|20 | ···················· | ···················· | ····················· | ···················· | ···················· | ···················· |
| S | 8790 hCoV-19/England/ALDP-B62A4F/2020\|EPI_ISL_656473\|20 | ···················· | ···················· | ····················· | ···················· | ···················· | ···················· |
| S | 8791 hCoV-19/England/ALDP-B622B0/2020\|EPI_ISL_656474\|20 | ···················· | ···················· | ····················· | ···················· | ···················· | ···················· |
| S | 8792 hCoV-19/England/ALDP-B63343/2020\|EPI_ISL_656475\|20 | ···················· | ···················· | ····················· | ···················· | ···················· | ···················· |
| S | 8793 hCoV-19/England/ALDP-B624E7/2020\|EPI_ISL_656476\|20 | ···················· | ···················· | ····················· | ···················· | ···················· | ···················· |
| S | 8794 hCoV-19/England/ALDP-B623BD/2020\|EPI_ISL_656477\|20 | ···················· | ···················· | ····················· | ···················· | ···················· | ···················· |
| S | 8795 hCoV-19/England/ALDP-B62AC7/2020\|EPI_ISL_656478\|20 | ···················· | ···················· | ····················· | ···················· | ···················· | ···················· |
| S | 8796 hCoV-19/England/ALDP-B61F6F/2020\|EPI_ISL_656479\|20 | ···················· | ···················· | ····················· | ···················· | ···················· | ···················· |
| S | 8797 hCoV-19/England/CAMC-B514FA/2020\|EPI_ISL_657747\|20 | ···················· | ···················· | ····················· | ···················· | ···················· | ···················· |
| S | 8798 hCoV-19/England/CAMC-B51CB6/2020\|EPI_ISL_657750\|20 | ···················· | ···················· | ····················· | ···················· | ······T············· | ···················· |
| S | 8799 hCoV-19/England/MILK-B55010/2020\|EPI_ISL_657751\|20 | ···················· | ···················· | ····················· | ···················· | ···················· | ···················· |
| S | 8800 hCoV-19/England/MILK-B5512C/2020\|EPI_ISL_657752\|20 | ···················· | ···················· | ····················· | ···················· | ···················· | ···················· |
| S | 8801 hCoV-19/England/MILK-B54E53/2020\|EPI_ISL_657753\|20 | ···················· | ···················· | ····················· | ···················· | ···················· | ···················· |
| S | 8802 hCoV-19/England/CAMC-B51DD1/2020\|EPI_ISL_657754\|20 | ···················· | ···················· | ····················· | ···················· | ···················· | ···················· |
| S | 8803 hCoV-19/England/MILK-B54DFC/2020\|EPI_ISL_657755\|20 | ···················· | ···················· | ····················· | ···················· | ···················· | ···················· |
| S | 8804 hCoV-19/England/CAMC-B51542/2020\|EPI_ISL_657756\|20 | ···················· | ···················· | ····················· | ···················· | ······T············· | ···················· |
| S | 8805 hCoV-19/England/MILK-B54E26/2020\|EPI_ISL_657757\|20 | ···················· | ···················· | ····················· | ···················· | ···················· | ···················· |
| S | 8806 hCoV-19/England/MILK-B54D47/2020\|EPI_ISL_657758\|20 | ···················· | ···················· | ····················· | ···················· | ···················· | ···················· |
| S | 8807 hCoV-19/England/CAMC-B51612/2020\|EPI_ISL_657759\|20 | ···················· | ···················· | ····················· | ···················· | ···················· | ···················· |
| S | 8808 hCoV-19/England/CAMC-B51560/2020\|EPI_ISL_657762\|20 | ···················· | ·A·················· | ····················· | ···················· | ···················· | ···A················ |
| S | 8809 hCoV-19/England/CAMC-B513FD/2020\|EPI_ISL_657763\|20 | ···················· | ···················· | ····················· | ···················· | ···················· | ···················· |
| S | 8810 hCoV-19/England/CAMC-B51D86/2020\|EPI_ISL_657764\|20 | ···················· | ···················· | ····················· | ···················· | ······T············· | ···················· |
| S | 8811 hCoV-19/England/CAMC-B511C6/2020\|EPI_ISL_657765\|20 | ···················· | ···················· | ····················· | ···················· | ···················· | ···················· |
| S | 8812 hCoV-19/England/MILK-B54E44/2020\|EPI_ISL_657768\|20 | ···················· | ···················· | ····················· | ···················· | ···················· | ···················· |
| S | 8813 hCoV-19/England/MILK-B55380/2020\|EPI_ISL_657770\|20 | ···················· | ···················· | ····················· | ···················· | ···················· | ···················· |
| S | 8814 hCoV-19/England/CAMC-B513DF/2020\|EPI_ISL_657771\|20 | ···················· | ···················· | ····················· | ···················· | ···················· | ···················· |
| S | 8815 hCoV-19/England/MILK-B54A7D/2020\|EPI_ISL_657772\|20 | ···················· | ···················· | ····················· | ···················· | ···················· | ···················· |
| S | 8816 hCoV-19/England/MILK-B54C59/2020\|EPI_ISL_657773\|20 | ···················· | ···················· | ····················· | ···················· | ···················· | ···················· |
| S | 8817 hCoV-19/England/MILK-B54EEA/2020\|EPI_ISL_657774\|20 | ···················· | ···················· | ····················· | ···················· | ···················· | ···················· |
| S | 8818 hCoV-19/England/MILK-B5549C/2020\|EPI_ISL_657775\|20 | ···················· | ···················· | ····················· | ···················· | ···················· | ···················· |
| S | 8819 hCoV-19/England/MILK-B552B0/2020\|EPI_ISL_657776\|20 | ···················· | ···················· | ····················· | ···················· | ···················· | ···················· |
| S | 8820 hCoV-19/England/MILK-B55520/2020\|EPI_ISL_657777\|20 | ···················· | ···················· | ····················· | ···················· | ···················· | ···················· |
| S | 8821 hCoV-19/England/MILK-B553AE/2020\|EPI_ISL_657779\|20 | ···················· | ···················· | ····················· | ···················· | ···················· | ···················· |
| S | 8822 hCoV-19/England/CAMC-B512C3/2020\|EPI_ISL_657780\|20 | ···················· | ···················· | ····················· | ···················· | ···················· | ···················· |
| S | 8823 hCoV-19/England/MILK-B54C4A/2020\|EPI_ISL_657781\|20 | ···················· | ···················· | ····················· | ···················· | ······T············· | ···················· |
| S | 8824 hCoV-19/England/CAMC-B51CD4/2020\|EPI_ISL_657782\|20 | ···················· | ···················· | ····················· | ···················· | ···················· | ···················· |
| S | 8825 hCoV-19/England/MILK-B5513B/2020\|EPI_ISL_657783\|20 | ···················· | ···················· | ····················· | ···················· | ···················· | ···················· |
| S | 8826 hCoV-19/England/MILK-B551E0/2020\|EPI_ISL_657785\|20 | ···················· | ···················· | ····················· | ···················· | ···················· | ···················· |
| S | 8827 hCoV-19/England/CAMC-B5122D/2020\|EPI_ISL_657786\|20 | ···················· | ·A·················· | ····················· | ···················· | ···················· | ···A················ |
| S | 8828 hCoV-19/England/CAMC-B514CD/2020\|EPI_ISL_657787\|20 | ···················· | ···················· | ····················· | ···················· | ···················· | ···················· |
| S | 8829 hCoV-19/England/MILK-B55274/2020\|EPI_ISL_657788\|20 | ···················· | ···················· | ····················· | ···················· | ···················· | ···················· |
| S | 8830 hCoV-19/England/MILK-B5520B/2020\|EPI_ISL_657789\|20 | ···················· | ·A·················· | ····················· | ···················· | ···················· | ···A················ |
| S | 8831 hCoV-19/England/MILK-B552CF/2020\|EPI_ISL_657791\|20 | ···················· | ···················· | ····················· | ···················· | ···················· | ···················· |
| S | 8832 hCoV-19/England/MILK-B54FE7/2020\|EPI_ISL_657792\|20 | ···················· | ···················· | ····················· | ···················· | ···················· | ···················· |
| S | 8833 hCoV-19/England/CAMC-B52101/2020\|EPI_ISL_657793\|20 | ···················· | ···················· | ····················· | ···················· | ······T············· | ···················· |
| S | 8834 hCoV-19/England/MILK-B54DB0/2020\|EPI_ISL_657794\|20 | ···················· | ·A·················· | ····················· | ···················· | ···················· | ···A················ |
| S | 8835 hCoV-19/England/MILK-B54CB3/2020\|EPI_ISL_657796\|20 | ···················· | ···················· | ····················· | ···················· | ···················· | ···················· |
| S | 8836 hCoV-19/England/MILK-B54C95/2020\|EPI_ISL_657797\|20 | ···················· | ···················· | ····················· | ···················· | ···················· | ···················· |
| T | 1 hCoV-19/England/QEUH-B809E4/2020\|EPI_ISL_661720\|20 | ···················· | ···················· | ····················· | ···················· | ···················· | ···················· |

```
T    2 hCoV-19/England/QEUH-B80559/2020|EPI_ISL_661722|20   ···················· ···················· ···················· ···················· ···················· ····················
T    3 hCoV-19/England/QEUH-B7FE61/2020|EPI_ISL_661723|20   ···················· ···················· ···················· ···················· ···················· ····················
T    4 hCoV-19/England/QEUH-B8234E/2020|EPI_ISL_661724|20   ···················· ·A·················· ···················· ···················· ···················· ···A················
T    5 hCoV-19/England/QEUH-B8229C/2020|EPI_ISL_661725|20   ···················· ···················· ···················· ···················· ···················· ····················
T    6 hCoV-19/England/CAMC-B7832D/2020|EPI_ISL_661726|20   ···················· ···················· ···················· ···················· ···················· ····················
T    7 hCoV-19/England/QEUH-B809F3/2020|EPI_ISL_661727|20   ···················· ···················· ···················· ···················· ···················· ····················
T    8 hCoV-19/England/QEUH-B80B66/2020|EPI_ISL_661730|20   ···················· ···················· ···················· ···················· ···················· ····················
T    9 hCoV-19/England/QEUH-B7FEBC/2020|EPI_ISL_661731|20   ···················· ···················· ···················· ···················· ···················· ····················
T   10 hCoV-19/England/QEUH-B80832/2020|EPI_ISL_661733|20   ···················· ···················· ···················· ···················· ···················· ····················
T   11 hCoV-19/England/CAMC-B77B44/2020|EPI_ISL_661734|20   ···················· ···················· ···················· ···················· ···················· ····················
T   12 hCoV-19/England/QEUH-B807BD/2020|EPI_ISL_661735|20   ···················· ···················· ···················· ···················· ···················· ····················
T   13 hCoV-19/England/QEUH-B7EFBA/2020|EPI_ISL_661736|20   ···················· ···················· ···················· ···················· ······T············· ····················
T   14 hCoV-19/England/QEUH-B80B1B/2020|EPI_ISL_661737|20   ···················· ·A·················· ···················· ···················· ···················· ···A················
T   15 hCoV-19/England/QEUH-B7F450/2020|EPI_ISL_661738|20   ···················· ···················· ···················· ···················· ···················· ····················
T   16 hCoV-19/England/QEUH-B7FF13/2020|EPI_ISL_661740|20   ···················· ···················· ···················· ···················· ···················· ····················
T   17 hCoV-19/England/CAMC-B77977/2020|EPI_ISL_661741|20   ···················· ···················· ···················· ···················· ······T············· ····················
T   18 hCoV-19/England/CAMC-B78545/2020|EPI_ISL_661742|20   ···················· ···················· ···················· ···················· ······T············· ····················
T   19 hCoV-19/England/QEUH-B7F098/2020|EPI_ISL_661744|20   ···················· ···················· ···················· ···················· ···················· ····················
T   20 hCoV-19/England/QEUH-B7FE43/2020|EPI_ISL_661745|20   ···················· ···················· ···················· ···················· ···················· ····················
T   21 hCoV-19/England/QEUH-B7EC95/2020|EPI_ISL_661746|20   ···················· ···················· ···················· ···················· ···················· ····················
T   22 hCoV-19/England/QEUH-B80A96/2020|EPI_ISL_661748|20   ···················· ···················· ···················· ···················· ···················· ····················
T   23 hCoV-19/England/CAMC-B78590/2020|EPI_ISL_661749|20   ···················· ···················· ···················· ···················· ···················· ····················
T   24 hCoV-19/England/CAMC-B77555/2020|EPI_ISL_661751|20   ···················· ···················· ···················· ···················· ···················· ····················
T   25 hCoV-19/England/QEUH-B7E3AF/2020|EPI_ISL_661753|20   ···················· ···················· ···················· ···················· ···················· ····················
T   26 hCoV-19/England/QEUH-B8228D/2020|EPI_ISL_661754|20   ···················· ·A·················· ···················· ···················· ···················· ···A················
T   27 hCoV-19/England/QEUH-B7FEDA/2020|EPI_ISL_661755|20   ···················· ···················· ···················· ···················· ···················· ····················
T   28 hCoV-19/England/QEUH-B82733/2020|EPI_ISL_661756|20   ···················· ···················· ···················· ···················· ···················· ····················
T   29 hCoV-19/England/QEUH-B7EF41/2020|EPI_ISL_661757|20   ···················· ···················· ···················· ···················· ···················· ····················
T   30 hCoV-19/England/QEUH-B7F432/2020|EPI_ISL_661758|20   ···················· ···················· ···················· ···················· ···················· ····················
T   31 hCoV-19/England/QEUH-B7ECA4/2020|EPI_ISL_661760|20   ···················· ···················· ···················· ···················· ···················· ····················
T   32 hCoV-19/England/CAMC-B78563/2020|EPI_ISL_661761|20   ···················· ···················· ···················· ···················· ···················· ····················
T   33 hCoV-19/England/QEUH-B82627/2020|EPI_ISL_661762|20   ···················· ···················· ···················· ···················· ···················· ····················
T   34 hCoV-19/England/QEUH-B82821/2020|EPI_ISL_661764|20   ···················· ·A·················· ···················· ···················· ···················· ····················
T   35 hCoV-19/England/QEUH-B7F423/2020|EPI_ISL_661765|20   ···················· ···················· ···················· ···················· ···················· ····················
T   36 hCoV-19/England/QEUH-B81084/2020|EPI_ISL_661766|20   ···················· ···················· ···················· ···················· ···················· ····················
T   37 hCoV-19/England/QEUH-B7EF32/2020|EPI_ISL_661767|20   ···················· ···················· ···················· ···················· ···················· ····················
T   38 hCoV-19/England/QEUH-B7FEF8/2020|EPI_ISL_661768|20   ···················· ···················· ···················· ···················· ···················· ····················
T   39 hCoV-19/England/QEUH-B7F010/2020|EPI_ISL_661770|20   ···················· ···················· ···················· ···················· ···················· ····················
T   40 hCoV-19/England/QEUH-B80577/2020|EPI_ISL_661771|20   ···················· ···················· ···················· ···················· ···················· ····················
T   41 hCoV-19/England/QEUH-B828B8/2020|EPI_ISL_661772|20   ···················· ···················· ···················· ···················· ···················· ····················
T   42 hCoV-19/England/QEUH-B805E0/2020|EPI_ISL_661773|20   ···················· ·A·················· ···················· ···················· ···················· ····················
T   43 hCoV-19/England/CAMC-B78396/2020|EPI_ISL_661775|20   ···················· ···················· ···················· ···················· ···················· ····················
T   44 hCoV-19/England/QEUH-B7EFAB/2020|EPI_ISL_661776|20   ···················· ···················· ···················· ···················· ···················· ····················
T   45 hCoV-19/England/QEUH-B807CC/2020|EPI_ISL_661777|20   ···················· ···················· ···················· ···················· ···················· ····················
T   46 hCoV-19/England/CAMC-B778F2/2020|EPI_ISL_661778|20   ···················· ···················· ···················· ···················· ······T············· ····················
T   47 hCoV-19/England/QEUH-B8053B/2020|EPI_ISL_661780|20   ···················· ···················· ···················· ···················· ···················· ····················
T   48 hCoV-19/England/CAMC-B77582/2020|EPI_ISL_661781|20   ···················· ···················· ···················· ···················· ···················· ····················
T   49 hCoV-19/England/QEUH-B80498/2020|EPI_ISL_661782|20   ···················· ···················· ···················· ···················· ···················· ····················
T   50 hCoV-19/England/CAMC-B77B53/2020|EPI_ISL_661783|20   ···················· ···················· ···················· ···················· ···················· ····················
T   51 hCoV-19/England/QEUH-B7ECC2/2020|EPI_ISL_661784|20   ···················· ···················· ···················· ···················· ···················· ····················
T   52 hCoV-19/England/QEUH-B7EF6F/2020|EPI_ISL_661786|20   ···················· ···················· ···················· ···················· ···················· ····················
T   53 hCoV-19/England/QEUH-B7FF6E/2020|EPI_ISL_661787|20   ···················· ···················· ···················· ···················· ···················· ····················
T   54 hCoV-19/England/QEUH-B7FE07/2020|EPI_ISL_661788|20   ···················· ···················· ···················· ···················· ···················· ····················
T   55 hCoV-19/England/QEUH-B7ECE0/2020|EPI_ISL_661790|20   ···················· ···················· ···················· ···················· ···················· ····················
T   56 hCoV-19/England/CAMC-B778C5/2020|EPI_ISL_661791|20   ···················· ···················· ···················· ···················· ···················· ····················
T   57 hCoV-19/England/QEUH-B7ED65/2020|EPI_ISL_661792|20   ···················· ···················· ···················· ···················· ···················· ····················
T   58 hCoV-19/England/QEUH-B80A0F/2020|EPI_ISL_661795|20   ···················· ···················· ···················· ···················· ···················· ····················
T   59 hCoV-19/England/QEUH-B7EFF6/2020|EPI_ISL_661798|20   ···················· ···················· ···················· ···················· ···················· ····················
T   60 hCoV-19/England/CAMC-B778D4/2020|EPI_ISL_661799|20   ···················· ···················· ···················· ···················· ···················· ····················
T   61 hCoV-19/Norway/CS-22/2020|EPI_ISL_666602|2020-03-2   ···················· ···················· ···················· ···················· ···················· ····················
T   62 hCoV-19/Norway/CS-24/2020|EPI_ISL_666604|2020-04-0   ···················· ·············A······ ···················· ···················· ···················· ····················
T   63 hCoV-19/Norway/CS-26/2020|EPI_ISL_666606|2020-04-0   ···················· ···················· ···················· ···················· ···················· ····················
T   64 hCoV-19/USA/LA-EVTL728/2020|EPI_ISL_666609|2020-07   ···················· ·············A······ ···················· ···················· ···················· ····················
T   65 hCoV-19/Norway/CS-29/2020|EPI_ISL_666610|2020-04-2   ···················· ···················· ···················· ···················· ···················· ····················
T   66 hCoV-19/Norway/CS-31/2020|EPI_ISL_666612|2020-04-2   ···················· ···················· ···················· ···················· ···················· ····················
T   67 hCoV-19/England/QEUH-B7E5F4/2020|EPI_ISL_667900|20   ···················· ···················· ···················· ···················· ···················· ····················
T   68 hCoV-19/England/CAMC-B79DA6/2020|EPI_ISL_667901|20   ···················· ···················· ···················· ···················· ···················· ····················
T   69 hCoV-19/England/CAMC-B7A1C7/2020|EPI_ISL_667902|20   ···················· ···················· ···················· ···················· ···················· ····················
T   70 hCoV-19/England/QEUH-B7E4F7/2020|EPI_ISL_667904|20   ···················· ···················· ···················· ···················· ···················· ····················
T   71 hCoV-19/England/CAMC-B799DF/2020|EPI_ISL_667905|20   ···················· ···················· ···················· ···················· ···················· ····················
T   72 hCoV-19/England/QEUH-B816DD/2020|EPI_ISL_667907|20   ···················· ···················· ···················· ···················· ···················· ····················
T   73 hCoV-19/England/CAMC-B7AE75/2020|EPI_ISL_667909|20   ···················· ···················· ···················· ···················· ···················· ····················
T   74 hCoV-19/England/CAMC-B7AE66/2020|EPI_ISL_667910|20   ···················· ···················· ···················· ···················· ···················· ····················
T   75 hCoV-19/England/CAMC-B78F56/2020|EPI_ISL_667911|20   ···················· ···················· ···················· ···················· ···················· ····················
T   76 hCoV-19/England/QEUH-B81716/2020|EPI_ISL_667912|20   ···················· ···················· ···················· ···················· ···················· ····················
T   77 hCoV-19/England/CAMC-B7AE84/2020|EPI_ISL_667913|20   ···················· ·A·················· ···················· ···················· ···················· ···A················
T   78 hCoV-19/England/QEUH-B81AFF/2020|EPI_ISL_667915|20   ···················· ···················· ···················· ···················· ···················· ····················
T   79 hCoV-19/England/QEUH-B7EDA1/2020|EPI_ISL_667917|20   ···················· ···················· ···················· ···················· ···················· ····················
T   80 hCoV-19/England/QEUH-B7EA13/2020|EPI_ISL_667918|20   ···················· ···················· ···················· ···················· ···················· ····················
T   81 hCoV-19/England/QEUH-B7EBB6/2020|EPI_ISL_667919|20   ···················· ···················· ···················· ···················· ···················· ····················
```

| | | | | | | | |
|---|---|---|---|---|---|---|---|
| T | 82 hCoV-19/England/CAMC-B797F3/2020\|EPI_ISL_667921\|20 | ···················· | ···················· | ···················· | ···················· | ···················· | ···················· |
| T | 83 hCoV-19/England/QEUH-B7E8FB/2020\|EPI_ISL_667922\|20 | ···················· | ···················· | ···················· | ···················· | ···················· | ···················· |
| T | 84 hCoV-19/England/CAMC-B78AA0/2020\|EPI_ISL_667923\|20 | ···················· | ···················· | ···················· | ···················· | ···················· | ···················· |
| T | 85 hCoV-19/England/QEUH-B81752/2020\|EPI_ISL_667924\|20 | ···················· | ···················· | ···················· | ···················· | ···················· | ···················· |
| T | 86 hCoV-19/England/QEUH-B7E9AD/2020\|EPI_ISL_667927\|20 | ···················· | ···················· | ···················· | ···················· | ···················· | ···················· |
| T | 87 hCoV-19/England/QEUH-B7EAC8/2020\|EPI_ISL_667928\|20 | ···················· | ···················· | ···················· | ···················· | ···················· | ···················· |
| T | 88 hCoV-19/England/QEUH-B7E4BB/2020\|EPI_ISL_667929\|20 | ···················· | ···················· | ···················· | ···················· | ···················· | ···················· |
| T | 89 hCoV-19/England/QEUH-B7EDB0/2020\|EPI_ISL_667931\|20 | ···················· | ·A·················· | ···················· | ···················· | ···················· | ···················· |
| T | 90 hCoV-19/England/CAMC-B79526/2020\|EPI_ISL_667933\|20 | ···················· | ···················· | ···················· | ···················· | ···················· | ···················· |
| T | 91 hCoV-19/England/CAMC-B78E77/2020\|EPI_ISL_667935\|20 | ···················· | ···················· | ···················· | ···················· | ···················· | ···················· |
| T | 92 hCoV-19/England/CAMC-B78DB6/2020\|EPI_ISL_667936\|20 | ···················· | ···················· | ···················· | ···················· | ···················· | ···················· |
| T | 93 hCoV-19/England/CAMC-B79D97/2020\|EPI_ISL_667937\|20 | ···················· | ···················· | ···················· | ···················· | ···················· | ···················· |
| T | 94 hCoV-19/England/QEUH-B816EC/2020\|EPI_ISL_667938\|20 | ···················· | ···················· | ···················· | ···················· | ···················· | ···················· |
| T | 95 hCoV-19/England/QEUH-B7EE17/2020\|EPI_ISL_667941\|20 | ···················· | ···················· | ···················· | ···················· | ···················· | ···················· |
| T | 96 hCoV-19/England/CAMC-B78FCF/2020\|EPI_ISL_667942\|20 | ···················· | ···················· | ···················· | ···················· | ···················· | ···················· |
| T | 97 hCoV-19/Denmark/DCGC-11731/2020\|EPI_ISL_671100\|202 | ···················· | ···················· | ···················· | ···················· | ···················· | ···················· |
| T | 98 hCoV-19/Denmark/DCGC-11732/2020\|EPI_ISL_671101\|202 | ···················· | ···················· | ···················· | ···················· | ···················· | ···················· |
| T | 99 hCoV-19/Denmark/DCGC-11733/2020\|EPI_ISL_671102\|202 | ···················· | ···················· | ···················· | ···················· | ···················· | ···················· |
| T | 100 hCoV-19/Denmark/DCGC-11735/2020\|EPI_ISL_671103\|202 | ···················· | ···················· | ···················· | ···················· | ···················· | ···················· |
| T | 101 hCoV-19/Denmark/DCGC-11736/2020\|EPI_ISL_671104\|202 | ···················· | ···················· | ···················· | ···················· | ···················· | ···················· |
| T | 102 hCoV-19/Denmark/DCGC-11737/2020\|EPI_ISL_671105\|202 | ···················· | ···················· | ···················· | ···················· | ···················· | ···················· |
| T | 103 hCoV-19/Denmark/DCGC-11738/2020\|EPI_ISL_671106\|202 | ···················· | ···················· | ···················· | ···················· | ···················· | ···················· |
| T | 104 hCoV-19/Denmark/DCGC-11741/2020\|EPI_ISL_671107\|202 | ···················· | ···················· | ···················· | ···················· | ···················· | ···················· |
| T | 105 hCoV-19/Denmark/DCGC-11744/2020\|EPI_ISL_671108\|202 | ···················· | ···················· | ···················· | ···················· | ···················· | ···················· |
| T | 106 hCoV-19/Denmark/DCGC-11745/2020\|EPI_ISL_671109\|202 | ···················· | ···················· | ···················· | ···················· | ···················· | ···················· |
| T | 107 hCoV-19/Denmark/DCGC-11747/2020\|EPI_ISL_671110\|202 | ···················· | ···················· | ···················· | ···················· | ···················· | ···················· |
| T | 108 hCoV-19/Denmark/DCGC-11748/2020\|EPI_ISL_671111\|202 | ···················· | ···················· | ···················· | ···················· | ···················· | ···················· |
| T | 109 hCoV-19/Denmark/DCGC-11751/2020\|EPI_ISL_671112\|202 | ···················· | ···················· | ···················· | ···················· | ···················· | ···················· |
| T | 110 hCoV-19/Denmark/DCGC-11753/2020\|EPI_ISL_671113\|202 | ···················· | ···················· | ···················· | ···················· | ···················· | ···················· |
| T | 111 hCoV-19/Denmark/DCGC-11754/2020\|EPI_ISL_671114\|202 | ···················· | ···················· | ···················· | ···················· | ···················· | ···················· |
| T | 112 hCoV-19/Denmark/DCGC-11756/2020\|EPI_ISL_671115\|202 | ···················· | ···················· | ···················· | ···················· | ···················· | ···················· |
| T | 113 hCoV-19/Denmark/DCGC-11760/2020\|EPI_ISL_671116\|202 | ···················· | ···················· | ···················· | ···················· | ···················· | ···················· |
| T | 114 hCoV-19/Denmark/DCGC-11761/2020\|EPI_ISL_671117\|202 | ···················· | ···················· | ···················· | ···················· | ···················· | ···················· |
| T | 115 hCoV-19/Denmark/DCGC-11762/2020\|EPI_ISL_671118\|202 | ···················· | ···················· | ···················· | ···················· | ···················· | ···················· |
| T | 116 hCoV-19/Denmark/DCGC-11763/2020\|EPI_ISL_671119\|202 | ···················· | ···················· | ···················· | ···················· | ···················· | ···················· |
| T | 117 hCoV-19/Denmark/DCGC-11764/2020\|EPI_ISL_671120\|202 | ···················· | ···················· | ···················· | ···················· | ···················· | ···················· |
| T | 118 hCoV-19/Denmark/DCGC-11765/2020\|EPI_ISL_671121\|202 | ···················· | ···················· | ···················· | ···················· | ······T············· | ···················· |
| T | 119 hCoV-19/Denmark/DCGC-11766/2020\|EPI_ISL_671122\|202 | ···················· | ···················· | ···················· | ···················· | ···················· | ···················· |
| T | 120 hCoV-19/Denmark/DCGC-11787/2020\|EPI_ISL_671136\|202 | ···················· | ···················· | ···················· | ···················· | ···················· | ···················· |
| T | 121 hCoV-19/Denmark/DCGC-11790/2020\|EPI_ISL_671138\|202 | ···················· | ···················· | ···················· | ···················· | ···················· | ···················· |
| T | 122 hCoV-19/Denmark/DCGC-11791/2020\|EPI_ISL_671139\|202 | ···················· | ···················· | ···················· | ···················· | ···················· | ···················· |
| T | 123 hCoV-19/Denmark/DCGC-11800/2020\|EPI_ISL_671145\|202 | ···················· | ···················· | ···················· | ···················· | ···················· | ···················· |
| T | 124 hCoV-19/Denmark/DCGC-11801/2020\|EPI_ISL_671146\|202 | ···················· | ···················· | ···················· | ···················· | ···················· | ···················· |
| T | 125 hCoV-19/Denmark/DCGC-11803/2020\|EPI_ISL_671147\|202 | ···················· | ···················· | ···················· | ···················· | ···················· | ···················· |
| T | 126 hCoV-19/Denmark/DCGC-11809/2020\|EPI_ISL_671148\|202 | ···················· | ···················· | ···················· | ···················· | ···················· | ···················· |
| T | 127 hCoV-19/Denmark/DCGC-11811/2020\|EPI_ISL_671149\|202 | ···················· | ···················· | ···················· | ···················· | ···················· | ···················· |
| T | 128 hCoV-19/Denmark/DCGC-11812/2020\|EPI_ISL_671150\|202 | ···················· | ···················· | ···················· | ···················· | ···················· | ···················· |
| T | 129 hCoV-19/Denmark/DCGC-11817/2020\|EPI_ISL_671151\|202 | ···················· | ···················· | ···················· | ···················· | ···················· | ···················· |
| T | 130 hCoV-19/Denmark/DCGC-11818/2020\|EPI_ISL_671152\|202 | ···················· | ···················· | ···················· | ···················· | ···················· | ···················· |
| T | 131 hCoV-19/Denmark/DCGC-11820/2020\|EPI_ISL_671153\|202 | ···················· | ···················· | ···················· | ···················· | ···················· | ···················· |
| T | 132 hCoV-19/Denmark/DCGC-11821/2020\|EPI_ISL_671154\|202 | ···················· | ·A·················· | ···················· | ···················· | ···················· | ···················· |
| T | 133 hCoV-19/Denmark/DCGC-11822/2020\|EPI_ISL_671155\|202 | ···················· | ···················· | ···················· | ···················· | ···················· | ···················· |
| T | 134 hCoV-19/Denmark/DCGC-11823/2020\|EPI_ISL_671156\|202 | ···················· | ···················· | ···················· | ···················· | ···················· | ···················· |
| T | 135 hCoV-19/Denmark/DCGC-11824/2020\|EPI_ISL_671157\|202 | ···················· | ···················· | ···················· | ···················· | ···················· | ···················· |
| T | 136 hCoV-19/Denmark/DCGC-11825/2020\|EPI_ISL_671158\|202 | ···················· | ···················· | ···················· | ···················· | ···················· | ···················· |
| T | 137 hCoV-19/Denmark/DCGC-11828/2020\|EPI_ISL_671159\|202 | ···················· | ···················· | ···················· | ···················· | ···················· | ···················· |
| T | 138 hCoV-19/Denmark/DCGC-11829/2020\|EPI_ISL_671160\|202 | ···················· | ···················· | ···················· | ···················· | ···················· | ···················· |
| T | 139 hCoV-19/Denmark/DCGC-11831/2020\|EPI_ISL_671161\|202 | ···················· | ···················· | ···················· | ···················· | ···················· | ···················· |
| T | 140 hCoV-19/Denmark/DCGC-11832/2020\|EPI_ISL_671162\|202 | ···················· | ···················· | ···················· | ···················· | ···················· | ···················· |
| T | 141 hCoV-19/Denmark/DCGC-11835/2020\|EPI_ISL_671163\|202 | ···················· | ···················· | ···················· | ···················· | ···················· | ···················· |
| T | 142 hCoV-19/Denmark/DCGC-11836/2020\|EPI_ISL_671164\|202 | ···················· | ·A·················· | ···················· | ···················· | ···················· | ···················· |
| T | 143 hCoV-19/Denmark/DCGC-11838/2020\|EPI_ISL_671165\|202 | ···················· | ···················· | ···················· | ···················· | ···················· | ···················· |
| T | 144 hCoV-19/Denmark/DCGC-11839/2020\|EPI_ISL_671166\|202 | ···················· | ·A·················· | ···················· | ···················· | ···················· | ···A················ |
| T | 145 hCoV-19/Denmark/DCGC-11841/2020\|EPI_ISL_671167\|202 | ···················· | ···················· | ···················· | ···················· | ···················· | ···················· |
| T | 146 hCoV-19/Denmark/DCGC-11842/2020\|EPI_ISL_671168\|202 | ···················· | ···················· | ···················· | ···················· | ···················· | ···················· |
| T | 147 hCoV-19/Denmark/DCGC-11843/2020\|EPI_ISL_671169\|202 | ···················· | ···················· | ···················· | ···················· | ···················· | ···················· |
| T | 148 hCoV-19/Denmark/DCGC-11844/2020\|EPI_ISL_671170\|202 | ···················· | ···················· | ···················· | ···················· | ···················· | ···················· |
| T | 149 hCoV-19/Denmark/DCGC-11845/2020\|EPI_ISL_671171\|202 | ···················· | ···················· | ···················· | ···················· | ···················· | ···················· |
| T | 150 hCoV-19/Denmark/DCGC-11846/2020\|EPI_ISL_671172\|202 | ···················· | ···················· | ···················· | ···················· | ···················· | ···················· |
| T | 151 hCoV-19/Denmark/DCGC-11847/2020\|EPI_ISL_671173\|202 | ···················· | ·A·················· | ···················· | ···················· | ···················· | ···A················ |
| T | 152 hCoV-19/Denmark/DCGC-11849/2020\|EPI_ISL_671174\|202 | ···················· | ···················· | ···················· | ···················· | ···················· | ···················· |
| T | 153 hCoV-19/Denmark/DCGC-11850/2020\|EPI_ISL_671175\|202 | ···················· | ·A·················· | ···················· | ···················· | ···················· | ···················· |
| T | 154 hCoV-19/Denmark/DCGC-11851/2020\|EPI_ISL_671176\|202 | ···················· | ···················· | ···················· | ···················· | ···················· | ···················· |
| T | 155 hCoV-19/Denmark/DCGC-11852/2020\|EPI_ISL_671177\|202 | ···················· | ·A·················· | ···················· | ···················· | ···················· | ···A················ |
| T | 156 hCoV-19/Denmark/DCGC-11853/2020\|EPI_ISL_671178\|202 | ···················· | ·A·················· | ···················· | ···················· | ···················· | ···················· |
| T | 157 hCoV-19/Denmark/DCGC-11855/2020\|EPI_ISL_671179\|202 | ···················· | ···················· | ···················· | ···················· | ···················· | ···················· |
| T | 158 hCoV-19/Denmark/DCGC-11856/2020\|EPI_ISL_671180\|202 | ···················· | ···················· | ···················· | ···················· | ···················· | ···················· |
| T | 159 hCoV-19/Denmark/DCGC-11858/2020\|EPI_ISL_671181\|202 | ···················· | ···················· | ···················· | ···················· | ···················· | ···················· |
| T | 160 hCoV-19/Denmark/DCGC-11859/2020\|EPI_ISL_671182\|202 | ···················· | ···················· | ···················· | ···················· | ···················· | ···················· |
| T | 161 hCoV-19/Denmark/DCGC-11860/2020\|EPI_ISL_671183\|202 | ···················· | ···················· | ···················· | ···················· | ···················· | ···················· |

```
T    162 hCoV-19/Denmark/DCGC-11861/2020|EPI_ISL_671184|202   ····················  ·A··················  ····················  ····················  ····················  ····················
T    163 hCoV-19/Denmark/DCGC-11864/2020|EPI_ISL_671185|202   ····················  ····················  ····················  ····················  ····················  ····················
T    164 hCoV-19/Denmark/DCGC-11865/2020|EPI_ISL_671186|202   ····················  ····················  ····················  ····················  ····················  ····················
T    165 hCoV-19/Denmark/DCGC-11867/2020|EPI_ISL_671187|202   ····················  ····················  ····················  ····················  ····················  ····················
T    166 hCoV-19/Denmark/DCGC-11868/2020|EPI_ISL_671188|202   ····················  ····················  ····················  ····················  ····················  ····················
T    167 hCoV-19/Denmark/DCGC-11869/2020|EPI_ISL_671189|202   ····················  ····················  ····················  ····················  ····················  ····················
T    168 hCoV-19/Denmark/DCGC-11872/2020|EPI_ISL_671190|202   ····················  ····················  ····················  ····················  ····················  ····················
T    169 hCoV-19/Denmark/DCGC-11876/2020|EPI_ISL_671191|202   ····················  ····················  ····················  ····················  ····················  ····················
T    170 hCoV-19/Denmark/DCGC-11880/2020|EPI_ISL_671192|202   ····················  ····················  ····················  ····················  ····················  ····················
T    171 hCoV-19/Denmark/DCGC-11881/2020|EPI_ISL_671193|202   ····················  ····················  ····················  ····················  ····················  ····················
T    172 hCoV-19/Denmark/DCGC-11882/2020|EPI_ISL_671194|202   ····················  ····················  ····················  ····················  ····················  ····················
T    173 hCoV-19/Denmark/DCGC-11883/2020|EPI_ISL_671195|202   ····················  ····················  ····················  ····················  ····················  ····················
T    174 hCoV-19/Denmark/DCGC-11885/2020|EPI_ISL_671196|202   ····················  ····················  ····················  ····················  ····················  ····················
T    175 hCoV-19/Denmark/DCGC-11887/2020|EPI_ISL_671197|202   ····················  ·A··················  ····················  ····················  ····················  ····················
T    176 hCoV-19/Denmark/DCGC-11889/2020|EPI_ISL_671198|202   ····················  ····················  ····················  ····················  ····················  ····················
T    177 hCoV-19/Denmark/DCGC-11890/2020|EPI_ISL_671199|202   ····················  ····················  ····················  ····················  ····················  ····················
T    178 hCoV-19/USA/CA-CZB-13137/2020|EPI_ISL_672400|2020-   ····················  ·A··················  ····················  ····················  ····················  ····················
T    179 hCoV-19/USA/CA-CZB-13135/2020|EPI_ISL_672401|2020-   ····················  ·A··················  ····················  ····················  ····················  ····················
T    181 hCoV-19/USA/CA-CZB-13133/2020|EPI_ISL_672403|2020-   ····················  ·A··················  ····················  ····················  ····················  ····················
T    182 hCoV-19/USA/CA-CZB-13132/2020|EPI_ISL_672404|2020-   ····················  ·A··················  ····················  ····················  ····················  ····················
T    183 hCoV-19/USA/CA-CZB-13131/2020|EPI_ISL_672405|2020-   ····················  ·A··················  ····················  ····················  ····················  ····················
T    184 hCoV-19/USA/CA-CZB-13130/2020|EPI_ISL_672406|2020-   ····················  ·A··················  ····················  ····················  ····················  ····················
T    186 hCoV-19/USA/CA-CZB-13127/2020|EPI_ISL_672408|2020-   ····················  ·A··················  ····················  ····················  ····················  ····················
T    187 hCoV-19/USA/CA-CZB-13126/2020|EPI_ISL_672409|2020-   ····················  ·A··················  ····················  ····················  ····················  ····················
T    188 hCoV-19/USA/CA-CZB-13125/2020|EPI_ISL_672410|2020-   ····················  ·A··················  ····················  ····················  ····················  ····················
T    189 hCoV-19/USA/CA-CZB-13124/2020|EPI_ISL_672411|2020-   ····················  ····················  ····················  ····················  ····················  ····················
T    190 hCoV-19/USA/CA-CZB-13122/2020|EPI_ISL_672412|2020-   ····················  ····················  ····················  ····················  ····················  ····················
T    191 hCoV-19/USA/CA-CZB-13121/2020|EPI_ISL_672413|2020-   ····················  ·A··················  ····················  ····················  ····················  ····················
T    192 hCoV-19/USA/CA-CZB-13120/2020|EPI_ISL_672414|2020-   ····················  ····················  ····················  ····················  ····················  ····················
T    193 hCoV-19/USA/CA-CZB-13119/2020|EPI_ISL_672415|2020-   ····················  ····················  ····················  ····················  ····················  ····················
T    194 hCoV-19/USA/CA-CZB-13118/2020|EPI_ISL_672416|2020-   ····················  ····················  ····················  ····················  ····················  ····················
T    195 hCoV-19/USA/CA-CZB-13117/2020|EPI_ISL_672417|2020-   ····················  ····················  ····················  ····················  ····················  ····················
T    196 hCoV-19/USA/CA-CZB-13116/2020|EPI_ISL_672418|2020-   ····················  ····················  ····················  ····················  ····················  ····················
T    197 hCoV-19/USA/CA-CZB-13115/2020|EPI_ISL_672419|2020-   ····················  ·A··················  ····················  ····················  ····················  ····················
T    199 hCoV-19/USA/CA-CZB-13111/2020|EPI_ISL_672421|2020-   ····················  ·A··················  ····················  ····················  ····················  ····················
T    200 hCoV-19/USA/CA-CZB-13110/2020|EPI_ISL_672422|2020-   ····················  ·A··················  ····················  ····················  ····················  ····················
T    203 hCoV-19/USA/CA-CZB-12880/2020|EPI_ISL_672425|2020-   ····················  ·A··················  ····················  ····················  ····················  ····················
T    204 hCoV-19/USA/CA-CZB-13106/2020|EPI_ISL_672426|2020-   ····················  ·A··················  ····················  ····················  ····················  ····················
T    206 hCoV-19/USA/CA-CZB-13108/2020|EPI_ISL_672428|2020-   ····················  ·A··················  ····················  ····················  ····················  ····················
T    209 hCoV-19/USA/CA-CZB-13273/2020|EPI_ISL_672431|2020-   ····················  ·A··················  ····················  ····················  ····················  ····················
T    211 hCoV-19/USA/CA-CZB-13289/2020|EPI_ISL_672433|2020-   ····················  ·A··················  ····················  ····················  ····················  ····················
T    212 hCoV-19/USA/CA-CZB-13303/2020|EPI_ISL_672434|2020-   ····················  ·A··················  ····················  ····················  ····················  ····················
T    214 hCoV-19/USA/CA-CZB-13312/2020|EPI_ISL_672436|2020-   ····················  ·A··················  ····················  ····················  ····················  ····················
T    215 hCoV-19/USA/CA-CZB-13331/2020|EPI_ISL_672437|2020-   ····················  ·A··················  ····················  ····················  ····················  ····················
T    216 hCoV-19/USA/CA-CZB-13336/2020|EPI_ISL_672438|2020-   ····················  ····················  ····················  ····················  ····················  ····················
T    217 hCoV-19/USA/CA-CZB-13341/2020|EPI_ISL_672439|2020-   ····················  ····················  ····················  ····················  ····················  ····················
T    218 hCoV-19/USA/CA-CZB-13342/2020|EPI_ISL_672440|2020-   ····················  ····················  ····················  ····················  ····················  ····················
T    221 hCoV-19/USA/CA-CZB-13347/2020|EPI_ISL_672443|2020-   ····················  ····················  ····················  ····················  ····················  ····················
T    223 hCoV-19/USA/CA-CZB-13351/2020|EPI_ISL_672445|2020-   ····················  ····················  ····················  ····················  ····················  ····················
T    224 hCoV-19/USA/CA-CZB-13353/2020|EPI_ISL_672446|2020-   ····················  ····················  ····················  ····················  ····················  ····················
T    225 hCoV-19/USA/CA-CZB-13354/2020|EPI_ISL_672447|2020-   ····················  ····················  ····················  ····················  ····················  ····················
T    227 hCoV-19/USA/CA-CZB-13362/2020|EPI_ISL_672449|2020-   ····················  ····················  ····················  ····················  ····················  ····················
T    228 hCoV-19/USA/CA-CZB-13373/2020|EPI_ISL_672450|2020-   ····················  ····················  ····················  ····················  ····················  ····················
T    230 hCoV-19/USA/CA-CZB-13375/2020|EPI_ISL_672452|2020-   ····················  ····················  ····················  ····················  ····················  ····················
T    232 hCoV-19/USA/CA-CZB-13425/2020|EPI_ISL_672469|2020-   ····················  ·A··················  ····················  ····················  ····················  ····················
T    233 hCoV-19/USA/CA-CZB-13459/2020|EPI_ISL_672476|2020-   ····················  ····················  ····················  ····················  ····················  ····················
T    234 hCoV-19/USA/CA-CZB-13238/2020|EPI_ISL_672477|2020-   ····················  ·A··················  ····················  ····················  ····················  ····················
T    235 hCoV-19/USA/CA-CZB-13239/2020|EPI_ISL_672478|2020-   ····················  ·A··················  ····················  ····················  ····················  ····················
T    236 hCoV-19/USA/CA-CZB-13240/2020|EPI_ISL_672479|2020-   ····················  ·A··················  ····················  ····················  ····················  ····················
T    237 hCoV-19/USA/CA-CZB-13241/2020|EPI_ISL_672480|2020-   ····················  ····················  ····················  ····················  ····················  ····················
T    238 hCoV-19/USA/CA-CZB-13242/2020|EPI_ISL_672481|2020-   ····················  ····················  ····················  ····················  ····················  ····················
T    239 hCoV-19/USA/CA-CZB-13243/2020|EPI_ISL_672482|2020-   ····················  ····················  ····················  ····················  ····················  ····················
T    240 hCoV-19/USA/CA-CZB-13244/2020|EPI_ISL_672483|2020-   ····················  ····················  ····················  ····················  ····················  ····················
T    241 hCoV-19/USA/CA-CZB-13245/2020|EPI_ISL_672484|2020-   ····················  ····················  ····················  ····················  ····················  ····················
T    243 hCoV-19/USA/CA-CZB-13247/2020|EPI_ISL_672486|2020-   ····················  ····················  ····················  ····················  ····················  ····················
T    244 hCoV-19/USA/CA-CZB-13248/2020|EPI_ISL_672487|2020-   ····················  ····················  ····················  ····················  ····················  ····················
T    245 hCoV-19/USA/CA-CZB-13535/2020|EPI_ISL_672488|2020-   ····················  ·A··················  ····················  ····················  ····················  ····················
T    246 hCoV-19/USA/CA-CZB-13461/2020|EPI_ISL_672489|2020-   ····················  ·A··················  ····················  ····················  ····················  ····················
T    247 hCoV-19/USA/CA-CZB-13551/2020|EPI_ISL_672490|2020-   ····················  ····················  ····················  ····················  ····················  ····················
T    248 hCoV-19/USA/CA-CZB-13550/2020|EPI_ISL_672491|2020-   ····················  ····················  ····················  ····················  ····················  ····················
T    249 hCoV-19/USA/CA-CZB-13548/2020|EPI_ISL_672492|2020-   ····················  ····················  ····················  ····················  NN··················  ····················
T    251 hCoV-19/USA/CA-CZB-13546/2020|EPI_ISL_672494|2020-   ····················  ····················  ····················  ····················  ····················  ····················
T    252 hCoV-19/USA/CA-CZB-13544/2020|EPI_ISL_672495|2020-   ····················  ····················  ····················  ····················  ····················  ····················
T    253 hCoV-19/USA/CA-CZB-13543/2020|EPI_ISL_672496|2020-   ····················  ····················  ····················  ····················  ····················  ····················
T    254 hCoV-19/USA/CA-CZB-13542/2020|EPI_ISL_672497|2020-   ····················  ····················  ····················  ····················  ····················  ····················
T    255 hCoV-19/USA/CA-CZB-13541/2020|EPI_ISL_672498|2020-   ····················  ····················  ····················  ····················  ····················  ····················
T    256 hCoV-19/USA/CA-CZB-13538/2020|EPI_ISL_672499|2020-   ····················  ····················  ····················  ····················  ····················  ····················
T    257 hCoV-19/England/QEUH-B81330/2020|EPI_ISL_661700|20   ····················  ····················  ····················  ····················  ····················  ····················
T    258 hCoV-19/England/QEUH-B7ED0B/2020|EPI_ISL_661701|20   ····················  ····················  ····················  ····················  ····················  ····················
T    259 hCoV-19/England/QEUH-B81039/2020|EPI_ISL_661703|20   ····················  ····················  ····················  ····················  ····················  ····················
```

| | | | | | | | | |
|---|---|---|---|---|---|---|---|---|
| T | 260 hCoV-19/England/QEUH-B7F02F/2020\|EPI_ISL_661704\|20 | ···················· | ···················· | ···················· | ···················· | ···················· | ···················· | |
| T | 261 hCoV-19/England/QEUH-B810A2/2020\|EPI_ISL_661706\|20 | ···················· | ···················· | ···················· | ···················· | ···················· | ···················· | |
| T | 262 hCoV-19/England/QEUH-B7ED1A/2020\|EPI_ISL_661708\|20 | ···················· | ···················· | ···················· | ···················· | ···················· | ···················· | |
| T | 263 hCoV-19/England/QEUH-B80814/2020\|EPI_ISL_661709\|20 | ···················· | ···················· | ···················· | ···················· | ···················· | ···················· | |
| T | 264 hCoV-19/England/CAMC-B787E5/2020\|EPI_ISL_661710\|20 | ···················· | ·A·················· | ···················· | ···················· | ···················· | ··A················· | |
| T | 265 hCoV-19/England/CAMC-B7794A/2020\|EPI_ISL_661712\|20 | ···················· | ·A·················· | ···················· | ···················· | ···················· | ··A················· | |
| T | 266 hCoV-19/England/QEUH-B804C5/2020\|EPI_ISL_661715\|20 | ···················· | ···················· | ···················· | ···················· | ···················· | ···················· | |
| T | 267 hCoV-19/England/QEUH-B805C2/2020\|EPI_ISL_661716\|20 | ···················· | ···················· | ···················· | ···················· | ···················· | ···················· | |
| T | 268 hCoV-19/England/QEUH-B82320/2020\|EPI_ISL_661718\|20 | ···················· | ···················· | ···················· | ···················· | ···················· | ···················· | |
| T | 269 hCoV-19/England/QEUH-B8101B/2020\|EPI_ISL_661719\|20 | ···················· | ···················· | ···················· | ···················· | ···················· | ···················· | |
| T | 270 hCoV-19/England/LOND-12501AF/2020\|EPI_ISL_665200\|2 | ···················· | ···················· | ···················· | ···················· | ···················· | ···················· | |
| T | 271 hCoV-19/England/LOND-12501EB/2020\|EPI_ISL_665201\|2 | ···················· | ···················· | ···················· | ···················· | ··NN················ | ···················· | |
| T | 272 hCoV-19/England/LOND-12501FA/2020\|EPI_ISL_665202\|2 | ···················· | ···················· | ···················· | ···················· | ···················· | ···················· | |
| T | 273 hCoV-19/England/LOND-1250206/2020\|EPI_ISL_665203\|2 | ···················· | ···················· | ···················· | ···················· | ···················· | ···················· | |
| T | 274 hCoV-19/England/LOND-1250224/2020\|EPI_ISL_665204\|2 | ···················· | ·A·················· | ···················· | ···················· | ···················· | ··A················· | |
| T | 275 hCoV-19/England/LOND-1250233/2020\|EPI_ISL_665205\|2 | ···················· | ···················· | ···················· | ···················· | ···················· | ···················· | |
| T | 276 hCoV-19/England/LOND-125028E/2020\|EPI_ISL_665206\|2 | ···················· | ···················· | ···················· | ···················· | ···················· | ···················· | |
| T | 277 hCoV-19/England/LOND-1250303/2020\|EPI_ISL_665207\|2 | ···················· | ···················· | ···················· | ···················· | ···················· | ···················· | |
| T | 278 hCoV-19/England/LOND-125039A/2020\|EPI_ISL_665208\|2 | ···················· | ···················· | ···················· | ···················· | ···················· | ···················· | |
| T | 279 hCoV-19/England/LOND-12503D6/2020\|EPI_ISL_665209\|2 | ···················· | ···················· | ···················· | ···················· | ···················· | ···················· | |
| T | 280 hCoV-19/England/LOND-125042E/2020\|EPI_ISL_665210\|2 | ···················· | ···················· | ···················· | ···················· | ···················· | ···················· | |
| T | 281 hCoV-19/England/LOND-1250497/2020\|EPI_ISL_665211\|2 | ···················· | ···················· | ···················· | ···················· | ···················· | ···················· | |
| T | 282 hCoV-19/England/LOND-12504A6/2020\|EPI_ISL_665212\|2 | ···················· | ···················· | ···················· | ···················· | ···················· | ···················· | |
| T | 283 hCoV-19/England/LOND-12504C4/2020\|EPI_ISL_665213\|2 | ···················· | ···················· | ···················· | ···················· | ···················· | ···················· | |
| T | 286 hCoV-19/England/LOND-1250558/2020\|EPI_ISL_665216\|2 | ···················· | ···················· | ···················· | ···················· | ···················· | ···················· | |
| T | 287 hCoV-19/England/LOND-1250585/2020\|EPI_ISL_665217\|2 | ···················· | ···················· | ···················· | ···················· | ···················· | ···················· | |
| T | 288 hCoV-19/England/LOND-12505B2/2020\|EPI_ISL_665218\|2 | ···················· | ···················· | ···················· | ···················· | ···················· | ···················· | |
| T | 289 hCoV-19/England/LOND-12505C1/2020\|EPI_ISL_665219\|2 | ···················· | ···················· | ···················· | ···················· | ···················· | ···················· | |
| T | 290 hCoV-19/England/LOND-12505EF/2020\|EPI_ISL_665220\|2 | ···················· | ···················· | ···················· | ···················· | ···················· | ···················· | |
| T | 291 hCoV-19/England/LOND-12505FE/2020\|EPI_ISL_665221\|2 | ···················· | ·A·················· | ···················· | ···················· | ···················· | ··A················· | |
| T | 292 hCoV-19/England/LOND-125060A/2020\|EPI_ISL_665222\|2 | ···················· | ···················· | ···················· | ···················· | ···················· | ···················· | |
| T | 293 hCoV-19/England/LOND-1250646/2020\|EPI_ISL_665223\|2 | ···················· | ···················· | ···················· | ···················· | ···················· | ···················· | |
| T | 294 hCoV-19/England/LOND-1250664/2020\|EPI_ISL_665224\|2 | ···················· | ···················· | ···················· | ···················· | ···················· | ···················· | |
| T | 295 hCoV-19/England/LOND-12506A0/2020\|EPI_ISL_665225\|2 | ···················· | ·A·················· | ···················· | ···················· | ···················· | ··A················· | |
| T | 296 hCoV-19/England/LOND-12506BF/2020\|EPI_ISL_665226\|2 | ···················· | ···················· | ···················· | ···················· | ···················· | ···················· | |
| T | 297 hCoV-19/England/LOND-12506FB/2020\|EPI_ISL_665227\|2 | ···················· | ···················· | ···················· | ···················· | ···················· | ···················· | |
| T | 299 hCoV-19/England/LOND-1250734/2020\|EPI_ISL_665229\|2 | ···················· | ···················· | ···················· | ···················· | ···················· | ···················· | |
| T | 300 hCoV-19/England/LOND-1250752/2020\|EPI_ISL_665230\|2 | ···················· | ···················· | ···················· | ···················· | ·····T·············· | ···················· | |
| T | 301 hCoV-19/England/LOND-1250761/2020\|EPI_ISL_665231\|2 | ···················· | ···················· | ···················· | ···················· | ···················· | ···················· | |
| T | 302 hCoV-19/England/LOND-12507AD/2020\|EPI_ISL_665232\|2 | ···················· | ···················· | ···················· | ···················· | ···················· | ···················· | |
| T | 303 hCoV-19/England/LOND-12507BC/2020\|EPI_ISL_665233\|2 | ···················· | ···················· | ···················· | ···················· | ···················· | ···················· | |
| T | 304 hCoV-19/England/LOND-12507CB/2020\|EPI_ISL_665234\|2 | ···················· | ···················· | ···················· | ···················· | ···················· | ···················· | |
| T | 305 hCoV-19/England/LOND-12507E9/2020\|EPI_ISL_665235\|2 | ···················· | ···················· | ···················· | ···················· | ···················· | ···················· | |
| T | 306 hCoV-19/England/LOND-125085F/2020\|EPI_ISL_665236\|2 | ···················· | ···················· | ···················· | ···················· | ···················· | ···················· | |
| T | 309 hCoV-19/England/LOND-12508F5/2020\|EPI_ISL_665239\|2 | ···················· | ···················· | ···················· | ···················· | ···················· | ···················· | |
| T | 310 hCoV-19/England/LOND-125093E/2020\|EPI_ISL_665240\|2 | ···················· | ···················· | ···················· | ···················· | ···················· | ···················· | |
| T | 311 hCoV-19/England/LOND-125096B/2020\|EPI_ISL_665241\|2 | ···················· | ···················· | ···················· | ···················· | ···················· | ···················· | |
| T | 312 hCoV-19/England/LOND-12509A7/2020\|EPI_ISL_665242\|2 | ···················· | ···················· | ···················· | ···················· | ···················· | ···················· | |
| T | 313 hCoV-19/England/LOND-1250AA4/2020\|EPI_ISL_665243\|2 | ···················· | ···················· | ···················· | ···················· | ···················· | ···················· | |
| T | 315 hCoV-19/England/LOND-1250AE0/2020\|EPI_ISL_665245\|2 | ···················· | ···················· | ···················· | ···················· | ···················· | ···················· | |
| T | 316 hCoV-19/England/LOND-1250AFF/2020\|EPI_ISL_665246\|2 | ···················· | ·A······A··········· | ···················· | ···················· | ···················· | ···················· | |
| T | 317 hCoV-19/England/LOND-1250B0B/2020\|EPI_ISL_665247\|2 | ···················· | ···················· | ···················· | ···················· | ·N··N··············· | ···················· | |
| T | 318 hCoV-19/England/LOND-1250B29/2020\|EPI_ISL_665248\|2 | ···················· | ···················· | ···················· | ···················· | ·····T·············· | ···················· | |
| T | 319 hCoV-19/England/LOND-1250B56/2020\|EPI_ISL_665249\|2 | ···················· | ···················· | ···················· | ···················· | NNN················· | ···················· | 1 |
| T | 321 hCoV-19/England/LOND-1250B92/2020\|EPI_ISL_665251\|2 | ···················· | ···················· | ···················· | ···················· | ···················· | ···················· | |
| T | 322 hCoV-19/Wales/PHWC-48CA73/2020\|EPI_ISL_666500\|2020 | ···················· | ···················· | ···················· | ···················· | ···················· | ···················· | |
| T | 323 hCoV-19/Wales/PHWC-48CA82/2020\|EPI_ISL_666501\|2020 | ···················· | ···················· | ···················· | ···················· | ···················· | ···················· | |
| T | 324 hCoV-19/Wales/PHWC-48CAA0/2020\|EPI_ISL_666502\|2020 | ···················· | ···················· | ···················· | ···················· | ···················· | ···················· | |
| T | 326 hCoV-19/Wales/PHWC-48CADD/2020\|EPI_ISL_666504\|2020 | ···················· | ···················· | ···················· | ···················· | ···················· | ···················· | |
| T | 327 hCoV-19/Wales/PHWC-48CAEC/2020\|EPI_ISL_666505\|2020 | ···················· | ···················· | ···················· | ···················· | ···················· | ···················· | |
| T | 328 hCoV-19/Wales/PHWC-48CAFB/2020\|EPI_ISL_666506\|2020 | ···················· | ···················· | ···················· | ···················· | ···················· | ···················· | |
| T | 329 hCoV-19/Wales/PHWC-48CB07/2020\|EPI_ISL_666507\|2020 | ···················· | ···················· | ···················· | ···················· | ···················· | ···················· | |
| T | 330 hCoV-19/Wales/PHWC-48CB16/2020\|EPI_ISL_666508\|2020 | ···················· | ···················· | ···················· | ···················· | ···················· | ···················· | |
| T | 331 hCoV-19/Wales/PHWC-48CB25/2020\|EPI_ISL_666509\|2020 | ···················· | ···················· | ···················· | ···················· | ···················· | ···················· | |
| T | 332 hCoV-19/Wales/PHWC-48CB43/2020\|EPI_ISL_666510\|2020 | ···················· | ···················· | ···················· | ···················· | ···················· | ···················· | |
| T | 333 hCoV-19/Wales/PHWC-48CB52/2020\|EPI_ISL_666511\|2020 | ···················· | ···················· | ···················· | ···················· | ···················· | ···················· | |
| T | 334 hCoV-19/Wales/PHWC-48CB61/2020\|EPI_ISL_666512\|2020 | ···················· | ···················· | ···················· | ···················· | ···················· | ···················· | |
| T | 336 hCoV-19/Wales/PHWC-48CB9E/2020\|EPI_ISL_666514\|2020 | ···················· | ···················· | ···················· | ···················· | ···················· | ···················· | |
| T | 337 hCoV-19/Wales/PHWC-48CBAD/2020\|EPI_ISL_666515\|2020 | ···················· | ···················· | ···················· | ···················· | ···················· | ···················· | |
| T | 338 hCoV-19/Wales/PHWC-48CBBC/2020\|EPI_ISL_666516\|2020 | ···················· | ···················· | ···················· | ···················· | ···················· | ···················· | |
| T | 339 hCoV-19/Wales/PHWC-48CBCB/2020\|EPI_ISL_666517\|2020 | ···················· | ···················· | ···················· | ···················· | ···················· | ···················· | |
| T | 340 hCoV-19/Wales/PHWC-48CBDA/2020\|EPI_ISL_666518\|2020 | ···················· | ···················· | ···················· | ···················· | ···················· | ···················· | |
| T | 341 hCoV-19/Wales/PHWC-48CBF8/2020\|EPI_ISL_666519\|2020 | ···················· | ···················· | ···················· | ···················· | ···················· | ···················· | |
| T | 342 hCoV-19/Wales/PHWC-48CC04/2020\|EPI_ISL_666520\|2020 | ···················· | ···················· | ···················· | ···················· | ···················· | ···················· | |
| T | 343 hCoV-19/Wales/PHWC-48CC13/2020\|EPI_ISL_666521\|2020 | ···················· | ···················· | ···················· | ···················· | ···················· | ···················· | |
| T | 344 hCoV-19/Wales/PHWC-48CC22/2020\|EPI_ISL_666522\|2020 | ···················· | ···················· | ···················· | ···················· | ···················· | ···················· | |
| T | 345 hCoV-19/Wales/PHWC-48CC6E/2020\|EPI_ISL_666523\|2020 | ···················· | ···················· | ···················· | ···················· | ···················· | ···················· | |
| T | 346 hCoV-19/Wales/PHWC-48CC7D/2020\|EPI_ISL_666524\|2020 | ···················· | ···················· | ···················· | ···················· | ···················· | ···················· | |
| T | 347 hCoV-19/Wales/PHWC-48CC8C/2020\|EPI_ISL_666525\|2020 | ···················· | ···················· | ···················· | ···················· | ···················· | ···················· | |
| T | 348 hCoV-19/Wales/PHWC-48CC9B/2020\|EPI_ISL_666526\|2020 | ···················· | ···················· | ···················· | ···················· | ···················· | ···················· | |

| | | | | | | | | |
|---|---|---|---|---|---|---|---|---|
| T | 349 hCoV-19/Wales/PHWC-48CCB9/2020\|EPI_ISL_666527\|2020 | ···················· | ···················· | ····················· | ···················· | ···················· | ···················· | |
| T | 350 hCoV-19/Wales/PHWC-48CCD7/2020\|EPI_ISL_666528\|2020 | ···················· | ···················· | ····················· | ···················· | ···················· | ···················· | |
| T | 352 hCoV-19/Wales/PHWC-48CD01/2020\|EPI_ISL_666530\|2020 | ···················· | ···················· | ····················· | ···················· | ···················· | ···················· | |
| T | 353 hCoV-19/Wales/PHWC-48CD10/2020\|EPI_ISL_666531\|2020 | ···················· | ···················· | ····················· | ···················· | ···················· | ···················· | |
| T | 354 hCoV-19/Wales/PHWC-48CD2F/2020\|EPI_ISL_666532\|2020 | ···················· | ···················· | ····················· | ···················· | ···················· | ···················· | |
| T | 355 hCoV-19/Wales/PHWC-48CD3E/2020\|EPI_ISL_666533\|2020 | ···················· | ···················· | ····················· | ···················· | ···················· | ···················· | |
| T | 357 hCoV-19/Wales/PHWC-48CD5C/2020\|EPI_ISL_666535\|2020 | ···················· | ···················· | ····················· | ···················· | ···················· | ···················· | |
| T | 358 hCoV-19/Wales/PHWC-48CD6B/2020\|EPI_ISL_666536\|2020 | ···················· | ···················· | ····················· | ···················· | ···················· | ···················· | |
| T | 359 hCoV-19/Wales/PHWC-48CD7A/2020\|EPI_ISL_666537\|2020 | ···················· | ···················· | ····················· | ···················· | ···················· | ···················· | |
| T | 360 hCoV-19/Wales/PHWC-48CD98/2020\|EPI_ISL_666538\|2020 | ···················· | ···················· | ····················· | ···················· | ···················· | ···················· | |
| T | 361 hCoV-19/Wales/PHWC-48CDB6/2020\|EPI_ISL_666539\|2020 | ···················· | ···················· | ····················· | ···················· | ···················· | ···················· | |
| T | 362 hCoV-19/Wales/PHWC-48CDD4/2020\|EPI_ISL_666540\|2020 | ···················· | ···················· | ····················· | ···················· | ···················· | ···················· | |
| T | 363 hCoV-19/Wales/PHWC-48CDE3/2020\|EPI_ISL_666541\|2020 | ···················· | ···················· | ····················· | ···················· | ···················· | ···················· | |
| T | 364 hCoV-19/Wales/PHWC-48CDF2/2020\|EPI_ISL_666542\|2020 | ···················· | ···················· | ····················· | ···················· | NNN················· | ···················· | 1 |
| T | 365 hCoV-19/Wales/PHWC-48CE0E/2020\|EPI_ISL_666543\|2020 | ···················· | ···················· | ····················· | ···················· | ···················· | ···················· | |
| T | 366 hCoV-19/Wales/PHWC-48CE1D/2020\|EPI_ISL_666544\|2020 | ···················· | ···················· | ····················· | ···················· | ···················· | ···················· | |
| T | 367 hCoV-19/Wales/PHWC-48CE2C/2020\|EPI_ISL_666545\|2020 | ···················· | ···················· | ····················· | ···················· | ···················· | ···················· | |
| T | 368 hCoV-19/Wales/PHWC-48CE4A/2020\|EPI_ISL_666546\|2020 | ···················· | ···················· | ····················· | ···················· | ···················· | ···················· | |
| T | 369 hCoV-19/Wales/PHWC-48CE59/2020\|EPI_ISL_666547\|2020 | ···················· | ···················· | ····················· | ···················· | ···················· | ···················· | |
| T | 370 hCoV-19/Wales/PHWC-48CE77/2020\|EPI_ISL_666548\|2020 | ···················· | ···················· | ····················· | ···················· | ···················· | ···················· | |
| T | 371 hCoV-19/Wales/PHWC-48CE86/2020\|EPI_ISL_666549\|2020 | ···················· | ···················· | ····················· | ···················· | ···················· | ···················· | |
| T | 372 hCoV-19/Wales/PHWC-48CE95/2020\|EPI_ISL_666550\|2020 | ···················· | ···················· | ····················· | ···················· | ···················· | ···················· | |
| T | 373 hCoV-19/Wales/PHWC-48CEA4/2020\|EPI_ISL_666551\|2020 | ···················· | ···················· | ····················· | ···················· | ···················· | ···················· | |
| T | 374 hCoV-19/Wales/PHWC-48CEC2/2020\|EPI_ISL_666552\|2020 | ···················· | ···················· | ····················· | ···················· | ···················· | ···················· | |
| T | 375 hCoV-19/Wales/PHWC-48CEE0/2020\|EPI_ISL_666553\|2020 | ···················· | ···················· | ····················· | ···················· | ···················· | ···················· | |
| T | 376 hCoV-19/Wales/PHWC-48CF0B/2020\|EPI_ISL_666554\|2020 | ···················· | ···················· | ····················· | ···················· | ···················· | ···················· | |
| T | 377 hCoV-19/Wales/PHWC-48CF1A/2020\|EPI_ISL_666555\|2020 | ···················· | ···················· | ····················· | ···················· | ···················· | ···················· | |
| T | 378 hCoV-19/Wales/PHWC-48CF38/2020\|EPI_ISL_666556\|2020 | ···················· | ···················· | ····················· | ···················· | ···················· | ···················· | |
| T | 379 hCoV-19/Wales/PHWC-48CF47/2020\|EPI_ISL_666557\|2020 | ···················· | ···················· | ····················· | ···················· | ···················· | ···················· | |
| T | 380 hCoV-19/Wales/PHWC-48CF56/2020\|EPI_ISL_666558\|2020 | ···················· | ···················· | ····················· | ···················· | ···················· | ···················· | |
| T | 381 hCoV-19/Wales/PHWC-48CF65/2020\|EPI_ISL_666559\|2020 | ···················· | ···················· | ····················· | ···················· | ···················· | ···················· | |
| T | 382 hCoV-19/Wales/PHWC-48CF83/2020\|EPI_ISL_666560\|2020 | ···················· | ···················· | ····················· | ···················· | ···················· | ···················· | |
| T | 383 hCoV-19/Wales/PHWC-48CF92/2020\|EPI_ISL_666561\|2020 | ···················· | ···················· | ····················· | ···················· | ···················· | ···················· | |
| T | 384 hCoV-19/Wales/PHWC-48CFA1/2020\|EPI_ISL_666562\|2020 | ···················· | ···················· | ····················· | ···················· | ···················· | ···················· | |
| T | 385 hCoV-19/Wales/PHWC-48CFCF/2020\|EPI_ISL_666563\|2020 | ···················· | ···················· | ····················· | ···················· | ···················· | ···················· | |
| T | 386 hCoV-19/Wales/PHWC-48CFDE/2020\|EPI_ISL_666564\|2020 | ···················· | ···················· | ····················· | ···················· | ···················· | ···················· | |
| T | 387 hCoV-19/Wales/PHWC-48CFFC/2020\|EPI_ISL_666565\|2020 | ···················· | ···················· | ····················· | ···················· | ···················· | ···················· | |
| T | 388 hCoV-19/Wales/PHWC-48D007/2020\|EPI_ISL_666566\|2020 | ···················· | ···················· | ····················· | ···················· | ···················· | ···················· | |
| T | 389 hCoV-19/Wales/PHWC-48D016/2020\|EPI_ISL_666567\|2020 | ···················· | ···················· | ····················· | ···················· | ···················· | ···················· | |
| T | 390 hCoV-19/Wales/PHWC-48D025/2020\|EPI_ISL_666569\|2020 | ···················· | ···················· | ····················· | ···················· | ···················· | ···················· | |
| T | 391 hCoV-19/Wales/PHWC-48D034/2020\|EPI_ISL_666570\|2020 | ···················· | ···················· | ····················· | ···················· | ···················· | ···················· | |
| T | 392 hCoV-19/Wales/PHWC-48D043/2020\|EPI_ISL_666571\|2020 | ···················· | ···················· | ····················· | ···················· | ···················· | ···················· | |
| T | 393 hCoV-19/Wales/PHWC-48D052/2020\|EPI_ISL_666572\|2020 | ···················· | ···················· | ····················· | ···················· | ···················· | ···················· | |
| T | 394 hCoV-19/Wales/PHWC-48D061/2020\|EPI_ISL_666573\|2020 | ···················· | ···················· | ····················· | ···················· | ···················· | ···················· | |
| T | 395 hCoV-19/Wales/PHWC-48D070/2020\|EPI_ISL_666574\|2020 | ···················· | ···················· | ····················· | ···················· | ···················· | ···················· | |
| T | 396 hCoV-19/Wales/PHWC-48D08F/2020\|EPI_ISL_666575\|2020 | ···················· | ···················· | ····················· | ···················· | ···················· | ···················· | |
| T | 397 hCoV-19/Wales/PHWC-48D09E/2020\|EPI_ISL_666576\|2020 | ···················· | ···················· | ····················· | ···················· | ···················· | ···················· | |
| T | 398 hCoV-19/Wales/PHWC-48D0AD/2020\|EPI_ISL_666577\|2020 | ···················· | ···················· | ····················· | ···················· | ···················· | ···················· | |
| T | 399 hCoV-19/Wales/PHWC-48D0BC/2020\|EPI_ISL_666578\|2020 | ···················· | ···················· | ····················· | ···················· | ···················· | ···················· | |
| T | 400 hCoV-19/Wales/PHWC-48D0CB/2020\|EPI_ISL_666579\|2020 | ···················· | ···················· | ····················· | ···················· | ···················· | ···················· | |
| T | 401 hCoV-19/Wales/PHWC-48D0F8/2020\|EPI_ISL_666580\|2020 | ···················· | ···················· | ····················· | ···················· | ···················· | ···················· | |
| T | 402 hCoV-19/Wales/PHWC-48D104/2020\|EPI_ISL_666581\|2020 | ···················· | ···················· | ····················· | ···················· | ···················· | ···················· | |
| T | 403 hCoV-19/Wales/PHWC-48D113/2020\|EPI_ISL_666582\|2020 | ···················· | ···················· | ····················· | ···················· | ···················· | ···················· | |
| T | 404 hCoV-19/Wales/PHWC-48D1AA/2020\|EPI_ISL_666590\|2020 | ···················· | ···················· | ····················· | ···················· | ···················· | ···················· | |
| T | 405 hCoV-19/Wales/QEUH-A60C88/2020\|EPI_ISL_666591\|2020 | ···················· | ···················· | ····················· | ···················· | ···················· | ···················· | |
| T | 406 hCoV-19/Wales/QEUH-AF0D8A/2020\|EPI_ISL_666592\|2020 | ···················· | ···················· | ····················· | ···················· | ···················· | ···················· | |
| T | 407 hCoV-19/Norway/CS-13/2020\|EPI_ISL_666593\|2020-04-1 | ···················· | ···················· | ····················· | ···················· | ···················· | ···················· | |
| T | 408 hCoV-19/Norway/CS-14/2020\|EPI_ISL_666594\|2020-04-1 | ···················· | ···················· | ····················· | ···················· | ···················· | ···················· | |
| T | 409 hCoV-19/Norway/CS-15/2020\|EPI_ISL_666595\|2020-06-1 | ···················· | ···················· | ····················· | ···················· | ···················· | ···················· | |
| T | 410 hCoV-19/Norway/CS-18/2020\|EPI_ISL_666598\|2020-06-2 | ···················· | ···················· | ····················· | ···················· | ···················· | ···················· | |
| T | 411 hCoV-19/England/CAMC-B79D5B/2020\|EPI_ISL_667823\|20 | ···················· | ···················· | ····················· | ···················· | ···················· | ···················· | |
| T | 412 hCoV-19/England/QEUH-B81AD1/2020\|EPI_ISL_667824\|20 | ···················· | ···················· | ····················· | ···················· | ···················· | ···················· | |
| T | 413 hCoV-19/England/QEUH-B816BF/2020\|EPI_ISL_667825\|20 | ···················· | ···················· | ····················· | ···················· | ···················· | ···················· | |
| T | 414 hCoV-19/England/CAMC-B78AEC/2020\|EPI_ISL_667826\|20 | ···················· | ···················· | ····················· | ···················· | ···················· | ···················· | |
| T | 415 hCoV-19/England/CAMC-B799C0/2020\|EPI_ISL_667827\|20 | ···················· | ···················· | ····················· | ···················· | ······T············· | ···················· | |
| T | 416 hCoV-19/England/QEUH-B7EEBD/2020\|EPI_ISL_667828\|20 | ···················· | ···················· | ····················· | ···················· | ···················· | ···················· | |
| T | 417 hCoV-19/England/QEUH-B7E9DA/2020\|EPI_ISL_667832\|20 | ···················· | ···················· | ····················· | ···················· | ···················· | ···················· | |
| T | 418 hCoV-19/England/CAMC-B7925C/2020\|EPI_ISL_667835\|20 | ···················· | ···················· | ····················· | ···················· | ···················· | ···················· | |
| T | 419 hCoV-19/England/QEUH-B7F1A4/2020\|EPI_ISL_667836\|20 | ···················· | ···················· | ····················· | ···················· | ···················· | ···················· | |
| T | 420 hCoV-19/England/QEUH-B80EB8/2020\|EPI_ISL_667838\|20 | ···················· | ···················· | ····················· | ···················· | ···················· | ···················· | |
| T | 421 hCoV-19/England/CAMC-B7AE1B/2020\|EPI_ISL_667840\|20 | ···················· | ·A·················· | ····················· | ···················· | ···················· | ···················· | |
| T | 422 hCoV-19/England/CAMC-B79CF4/2020\|EPI_ISL_667842\|20 | ···················· | ···················· | ····················· | ···················· | ···················· | ···················· | |
| T | 423 hCoV-19/England/QEUH-B81682/2020\|EPI_ISL_667843\|20 | ···················· | ···················· | ····················· | ···················· | ···················· | ···················· | |
| T | 424 hCoV-19/England/QEUH-B7E961/2020\|EPI_ISL_667849\|20 | ···················· | ···················· | ····················· | ···················· | ···················· | ···················· | |
| T | 425 hCoV-19/England/QEUH-B81A77/2020\|EPI_ISL_667851\|20 | ···················· | ···················· | ····················· | ···················· | ···················· | ···················· | |
| T | 426 hCoV-19/England/CAMC-B79113/2020\|EPI_ISL_667852\|20 | ···················· | ···················· | ····················· | ···················· | ···················· | ···················· | |
| T | 427 hCoV-19/England/CAMC-B78A28/2020\|EPI_ISL_667853\|20 | ···················· | ···················· | ····················· | ···················· | ···················· | ···················· | |
| T | 428 hCoV-19/England/QEUH-B7E62E/2020\|EPI_ISL_667855\|20 | ···················· | ···················· | ····················· | ···················· | ···················· | ···················· | |
| T | 429 hCoV-19/England/CAMC-B78976/2020\|EPI_ISL_667856\|20 | ···················· | ···················· | ····················· | ···················· | ···················· | ···················· | |
| T | 430 hCoV-19/England/QEUH-B81A2C/2020\|EPI_ISL_667857\|20 | ···················· | ···················· | ····················· | ···················· | ···················· | ···················· | |

| | | | | | | | | | |
|---|---|---|---|---|---|---|---|---|---|
| T | 431 | hCoV-19/England/CAMC-B79A36/2020\|EPI_ISL_667858\|20 | ···················· | ···················· | ···················· | ···················· | ···················· | ···················· | |
| T | 432 | hCoV-19/England/QEUH-B7E99E/2020\|EPI_ISL_667859\|20 | ···················· | ·A·················· | ···················· | ···················· | ···················· | ···················· | |
| T | 433 | hCoV-19/England/QEUH-B80FC4/2020\|EPI_ISL_667860\|20 | ···················· | ···················· | ···················· | ···················· | ···················· | ···················· | |
| T | 434 | hCoV-19/England/CAMC-B79DB5/2020\|EPI_ISL_667862\|20 | ···················· | ···················· | ···················· | ···················· | ···················· | ···················· | |
| T | 435 | hCoV-19/England/QEUH-B7F13B/2020\|EPI_ISL_667863\|20 | ···················· | ···················· | ···················· | ···················· | ···················· | ···················· | |
| T | 436 | hCoV-19/England/CAMC-B79948/2020\|EPI_ISL_667865\|20 | ···················· | ···················· | ···················· | ···················· | ···················· | ···················· | |
| T | 437 | hCoV-19/England/QEUH-B7E600/2020\|EPI_ISL_667866\|20 | ···················· | ···················· | ···················· | ···················· | ···················· | ···················· | |
| T | 438 | hCoV-19/England/CAMC-B78DC5/2020\|EPI_ISL_667869\|20 | ···················· | ···················· | ···················· | ···················· | ···················· | ···················· | |
| T | 439 | hCoV-19/England/CAMC-B79D00/2020\|EPI_ISL_667870\|20 | ···················· | ···················· | ···················· | ···················· | ···················· | ···················· | |
| T | 440 | hCoV-19/England/CAMC-B79DF1/2020\|EPI_ISL_667871\|20 | ···················· | ···················· | ···················· | ···················· | ···················· | ···················· | |
| T | 441 | hCoV-19/England/QEUH-B81A1D/2020\|EPI_ISL_667872\|20 | ···················· | ···················· | ···················· | ···················· | ···················· | ···················· | |
| T | 442 | hCoV-19/England/QEUH-B7F14A/2020\|EPI_ISL_667873\|20 | ···················· | ···················· | ···················· | ···················· | ···················· | ···················· | |
| T | 443 | hCoV-19/England/CAMC-B790E9/2020\|EPI_ISL_667874\|20 | ···················· | ···················· | ···················· | ···················· | ···················· | ···················· | |
| T | 444 | hCoV-19/England/CAMC-B78F83/2020\|EPI_ISL_667875\|20 | ···················· | ···················· | ···················· | ···················· | ···················· | ···················· | |
| T | 445 | hCoV-19/England/CAMC-B796AB/2020\|EPI_ISL_667877\|20 | ···················· | ···················· | ···················· | ···················· | ···················· | ···················· | |
| T | 446 | hCoV-19/England/CAMC-B78ACE/2020\|EPI_ISL_667878\|20 | ···················· | ···················· | ···················· | ···················· | ···················· | ···················· | |
| T | 447 | hCoV-19/England/CAMC-B78967/2020\|EPI_ISL_667880\|20 | ···················· | ···················· | ···················· | ···················· | ···················· | ···················· | |
| T | 448 | hCoV-19/England/QEUH-B7E4D9/2020\|EPI_ISL_667881\|20 | ···················· | ···················· | ···················· | ···················· | ···················· | ···················· | |
| T | 449 | hCoV-19/England/QEUH-B81646/2020\|EPI_ISL_667883\|20 | ···················· | ···················· | ···················· | ···················· | ···················· | ···················· | |
| T | 450 | hCoV-19/England/QEUH-B7EEF9/2020\|EPI_ISL_667885\|20 | ···················· | ···················· | ···················· | ···················· | ···················· | ···················· | |
| T | 451 | hCoV-19/England/CAMC-B78FB0/2020\|EPI_ISL_667886\|20 | ···················· | ···················· | ···················· | ···················· | ···················· | ···················· | |
| T | 452 | hCoV-19/England/QEUH-B7EA22/2020\|EPI_ISL_667887\|20 | ···················· | ···················· | ···················· | ···················· | ···················· | ···················· | |
| T | 453 | hCoV-19/England/QEUH-B81734/2020\|EPI_ISL_667888\|20 | ···················· | ···················· | ···················· | ···················· | ···················· | ···················· | |
| T | 454 | hCoV-19/England/QEUH-B7E530/2020\|EPI_ISL_667889\|20 | ···················· | ···················· | ···················· | ···················· | ···················· | ···················· | |
| T | 455 | hCoV-19/England/CAMC-B79104/2020\|EPI_ISL_667890\|20 | ···················· | ···················· | ···················· | ···················· | ······T············· | ···················· | |
| T | 456 | hCoV-19/England/QEUH-B81691/2020\|EPI_ISL_667891\|20 | ···················· | ···················· | ···················· | ···················· | ···················· | ···················· | |
| T | 457 | hCoV-19/England/QEUH-B7EDED/2020\|EPI_ISL_667893\|20 | ···················· | ···················· | ···················· | ···················· | ···················· | ···················· | |
| T | 458 | hCoV-19/England/CAMC-B79201/2020\|EPI_ISL_667894\|20 | ···················· | ···················· | ···················· | ···················· | ······T············· | ···················· | |
| T | 459 | hCoV-19/England/QEUH-B81A0E/2020\|EPI_ISL_667895\|20 | ···················· | ···················· | ···················· | ···················· | ···················· | ···················· | |
| T | 460 | hCoV-19/England/CAMC-B79DE2/2020\|EPI_ISL_667896\|20 | ···················· | ···················· | ···················· | ···················· | ···················· | ···················· | |
| T | 461 | hCoV-19/England/CAMC-B78949/2020\|EPI_ISL_667897\|20 | ···················· | ·A·················· | ···················· | ···················· | ···················· | ···A················ | |
| T | 462 | hCoV-19/England/QEUH-B7EA6E/2020\|EPI_ISL_667898\|20 | ···················· | ···················· | ···················· | ···················· | ···················· | ···················· | |
| T | 463 | hCoV-19/Denmark/DCGC-11715/2020\|EPI_ISL_671090\|202 | ···················· | ···················· | ···················· | ···················· | ···················· | ···················· | |
| T | 464 | hCoV-19/Denmark/DCGC-11719/2020\|EPI_ISL_671091\|202 | ···················· | ···················· | ···················· | ···················· | ···················· | ···················· | |
| T | 465 | hCoV-19/Denmark/DCGC-11720/2020\|EPI_ISL_671092\|202 | ···················· | ···················· | ···················· | ···················· | ···················· | ···················· | |
| T | 466 | hCoV-19/Denmark/DCGC-11721/2020\|EPI_ISL_671093\|202 | ···················· | ···················· | ···················· | ···················· | ···················· | ···················· | |
| T | 467 | hCoV-19/Denmark/DCGC-11722/2020\|EPI_ISL_671094\|202 | ···················· | ···················· | ···················· | ···················· | ···················· | ···················· | |
| T | 468 | hCoV-19/Denmark/DCGC-11724/2020\|EPI_ISL_671095\|202 | ···················· | ···················· | ···················· | ···················· | ···················· | ···················· | |
| T | 469 | hCoV-19/Denmark/DCGC-11725/2020\|EPI_ISL_671096\|202 | ···················· | ·A·················· | ···················· | ···················· | ···················· | ···················· | |
| T | 470 | hCoV-19/Denmark/DCGC-11726/2020\|EPI_ISL_671097\|202 | ···················· | ···················· | ···················· | ···················· | ···················· | ···················· | |
| T | 471 | hCoV-19/Denmark/DCGC-11728/2020\|EPI_ISL_671098\|202 | ···················· | ···················· | ···················· | ···················· | ···················· | ···················· | |
| T | 472 | hCoV-19/Denmark/DCGC-11729/2020\|EPI_ISL_671099\|202 | ···················· | ···················· | ···················· | ···················· | ···················· | ···················· | |
| T | 473 | hCoV-19/Denmark/DCGC-11767/2020\|EPI_ISL_671123\|202 | ···················· | ···················· | ···················· | ···················· | ···················· | ···················· | |
| T | 474 | hCoV-19/Denmark/DCGC-11770/2020\|EPI_ISL_671124\|202 | ···················· | ·A·················· | ···················· | ···················· | ···················· | ···················· | |
| T | 475 | hCoV-19/Denmark/DCGC-11771/2020\|EPI_ISL_671125\|202 | ···················· | ···················· | ···················· | ···················· | ···················· | ···················· | |
| T | 476 | hCoV-19/Denmark/DCGC-11772/2020\|EPI_ISL_671126\|202 | ···················· | ···················· | ···················· | ···················· | ······T············· | ···················· | |
| T | 477 | hCoV-19/Denmark/DCGC-11773/2020\|EPI_ISL_671127\|202 | ···················· | ···················· | ···················· | ···················· | ···················· | ···················· | |
| T | 478 | hCoV-19/Denmark/DCGC-11774/2020\|EPI_ISL_671128\|202 | ···················· | ·A·················· | ···················· | ···················· | ···················· | ···················· | |
| T | 479 | hCoV-19/Denmark/DCGC-11775/2020\|EPI_ISL_671129\|202 | ···················· | ···················· | ···················· | ···················· | ···················· | ···················· | |
| T | 480 | hCoV-19/Denmark/DCGC-11776/2020\|EPI_ISL_671130\|202 | ···················· | ···················· | ···················· | ···················· | ···················· | ···················· | |
| T | 481 | hCoV-19/Denmark/DCGC-11779/2020\|EPI_ISL_671131\|202 | ···················· | ···················· | ···················· | ···················· | ···················· | ···················· | |
| T | 482 | hCoV-19/Denmark/DCGC-11780/2020\|EPI_ISL_671132\|202 | ···················· | ···················· | ···················· | ···················· | ···················· | ···················· | |
| T | 483 | hCoV-19/Denmark/DCGC-11782/2020\|EPI_ISL_671133\|202 | ···················· | ···················· | ···················· | ···················· | ···················· | ···················· | |
| T | 484 | hCoV-19/Denmark/DCGC-11784/2020\|EPI_ISL_671134\|202 | ···················· | ···················· | ···················· | ···················· | ···················· | ···················· | |
| T | 485 | hCoV-19/Denmark/DCGC-11785/2020\|EPI_ISL_671135\|202 | ········T··········· | ···················· | ···················· | ···················· | ···················· | ···················· | |
| T | 486 | hCoV-19/Denmark/DCGC-11788/2020\|EPI_ISL_671137\|202 | ···················· | ·A·················· | ···················· | ···················· | ···················· | ···················· | |
| T | 487 | hCoV-19/Denmark/DCGC-11792/2020\|EPI_ISL_671140\|202 | ···················· | ···················· | ···················· | ···················· | ···················· | ···················· | |
| T | 488 | hCoV-19/Denmark/DCGC-11794/2020\|EPI_ISL_671141\|202 | ···················· | ···················· | ···················· | ···················· | ···················· | ···················· | |
| T | 489 | hCoV-19/Denmark/DCGC-11795/2020\|EPI_ISL_671142\|202 | ···················· | ·A·················· | ···················· | ···················· | ···················· | ···················· | |
| T | 490 | hCoV-19/Denmark/DCGC-11796/2020\|EPI_ISL_671143\|202 | ···················· | ···················· | ···················· | ···················· | ···················· | ···················· | |
| T | 491 | hCoV-19/Denmark/DCGC-11797/2020\|EPI_ISL_671144\|202 | ···················· | ···················· | ···················· | ···················· | ······T············· | ···················· | |
| T | 492 | hCoV-19/USA/CA-CZB-13384/2020\|EPI_ISL_672454\|2020- | ···················· | ·A·················· | ···················· | ···················· | ···················· | ···················· | |
| T | 493 | hCoV-19/USA/CA-CZB-13388/2020\|EPI_ISL_672455\|2020- | ···················· | ·A·················· | ···················· | ···················· | ···················· | ···················· | |
| T | 495 | hCoV-19/USA/CA-CZB-13396/2020\|EPI_ISL_672457\|2020- | ···················· | ·A·················· | ···················· | ···················· | ···················· | ···················· | |
| T | 497 | hCoV-19/USA/CA-CZB-13401/2020\|EPI_ISL_672459\|2020- | ···················· | ···················· | ···················· | ···················· | NNN················· | ···················· | 1 |
| T | 500 | hCoV-19/USA/CA-CZB-13410/2020\|EPI_ISL_672462\|2020- | ···················· | ·A·················· | ···················· | ···················· | ···················· | ···················· | |
| T | 501 | hCoV-19/USA/CA-CZB-13413/2020\|EPI_ISL_672463\|2020- | ···················· | ···················· | ···················· | ···················· | ···················· | ···················· | |
| T | 502 | hCoV-19/USA/CA-CZB-13417/2020\|EPI_ISL_672464\|2020- | ···················· | ·A·················· | ···················· | ···················· | ···················· | ···················· | |
| T | 503 | hCoV-19/USA/CA-CZB-13418/2020\|EPI_ISL_672465\|2020- | ···················· | ·A·················· | ···················· | ···················· | ···················· | ···················· | |
| T | 504 | hCoV-19/USA/CA-CZB-13419/2020\|EPI_ISL_672466\|2020- | ···················· | ····A··············· | ···················· | ···················· | ···················· | ···················· | |
| T | 506 | hCoV-19/USA/CA-CZB-13424/2020\|EPI_ISL_672468\|2020- | ···················· | ···················· | ···················· | ···················· | ···················· | ···················· | |
| T | 507 | hCoV-19/USA/CA-CZB-13444/2020\|EPI_ISL_672470\|2020- | ···················· | ···················· | ···················· | ···················· | ···················· | ···················· | |
| T | 508 | hCoV-19/USA/CA-CZB-13448/2020\|EPI_ISL_672471\|2020- | ···················· | ·A·················· | ···················· | ···················· | ···················· | ···················· | |
| T | 509 | hCoV-19/USA/CA-CZB-13449/2020\|EPI_ISL_672472\|2020- | ···················· | ···················· | ···················· | ···················· | ···················· | ···················· | |
| T | 510 | hCoV-19/USA/CA-CZB-13452/2020\|EPI_ISL_672473\|2020- | ···················· | ···················· | ···················· | ···················· | ···················· | ···················· | |
| T | 512 | hCoV-19/USA/CA-CZB-13456/2020\|EPI_ISL_672475\|2020- | ···················· | ·A·················· | ···················· | ···················· | ···················· | ···················· | |
| T | 513 | hCoV-19/England/QEUH-B6C59F/2020\|EPI_ISL_661600\|20 | ···················· | ···················· | ···················· | ···················· | ···················· | ···················· | |
| T | 514 | hCoV-19/England/QEUH-B6C571/2020\|EPI_ISL_661602\|20 | ···················· | ···················· | ···················· | ···················· | ···················· | ···················· | |
| T | 515 | hCoV-19/England/QEUH-B67BFC/2020\|EPI_ISL_661603\|20 | ···················· | ···················· | ···················· | ···················· | ···················· | ···················· | |
| T | 516 | hCoV-19/England/QEUH-B698A8/2020\|EPI_ISL_661604\|20 | ···················· | ···················· | ···················· | ···················· | ···················· | ···················· | |

| | | | | | | | |
|---|---|---|---|---|---|---|---|
| T | 517 hCoV-19/England/QEUH-B68A3A/2020\|EPI_ISL_661605\|20 | ···················· | ···················· | ····················· | ···················· | ···················· | ···················· |
| T | 518 hCoV-19/England/QEUH-B69978/2020\|EPI_ISL_661607\|20 | ···················· | ···················· | ····················· | ···················· | ···················· | ···················· |
| T | 519 hCoV-19/England/QEUH-B67A1D/2020\|EPI_ISL_661608\|20 | ···················· | ···················· | ····················· | ···················· | ···················· | ···················· |
| T | 520 hCoV-19/England/QEUH-B67BDE/2020\|EPI_ISL_661609\|20 | ···················· | ···················· | ····················· | ···················· | ···················· | ···················· |
| T | 521 hCoV-19/England/QEUH-B6C5CC/2020\|EPI_ISL_661610\|20 | ···················· | ···················· | ····················· | ···················· | ···················· | ···················· |
| T | 522 hCoV-19/England/QEUH-B67716/2020\|EPI_ISL_661611\|20 | ···················· | ···················· | ····················· | ···················· | ···················· | ···················· |
| T | 523 hCoV-19/England/QEUH-B66665/2020\|EPI_ISL_661612\|20 | ···················· | ···················· | ····················· | ···················· | ···················· | ···················· |
| T | 524 hCoV-19/England/QEUH-B67F1E/2020\|EPI_ISL_661613\|20 | ···················· | ···················· | ····················· | ···················· | ···················· | ···················· |
| T | 525 hCoV-19/England/QEUH-B689B5/2020\|EPI_ISL_661614\|20 | ···················· | ···················· | ····················· | ···················· | ···················· | ···················· |
| T | 526 hCoV-19/England/QEUH-B67E8A/2020\|EPI_ISL_661615\|20 | ···················· | ···················· | ····················· | ···················· | ···················· | ···················· |
| T | 527 hCoV-19/England/QEUH-B6986C/2020\|EPI_ISL_661616\|20 | ···················· | ···················· | ····················· | ···················· | ···················· | ···················· |
| T | 528 hCoV-19/England/QEUH-B66322/2020\|EPI_ISL_661617\|20 | ···················· | ···················· | ····················· | ···················· | ···················· | ···················· |
| T | 529 hCoV-19/England/QEUH-B66638/2020\|EPI_ISL_661618\|20 | ···················· | ···················· | ····················· | ···················· | ···················· | ···················· |
| T | 530 hCoV-19/England/QEUH-B6629E/2020\|EPI_ISL_661620\|20 | ···················· | ···················· | ····················· | ···················· | ···················· | ···················· |
| T | 531 hCoV-19/England/QEUH-B6C526/2020\|EPI_ISL_661622\|20 | ···················· | ···················· | ····················· | ···················· | ···················· | ···················· |
| T | 532 hCoV-19/England/QEUH-B66340/2020\|EPI_ISL_661624\|20 | ···················· | ···················· | ····················· | ···················· | ···················· | ···················· |
| T | 533 hCoV-19/England/QEUH-B67989/2020\|EPI_ISL_661626\|20 | ···················· | ···················· | ····················· | ···················· | ···················· | ···················· |
| T | 534 hCoV-19/England/QEUH-B681EA/2020\|EPI_ISL_661627\|20 | ···················· | ···················· | ····················· | ···················· | ···················· | ···················· |
| T | 535 hCoV-19/England/QEUH-B67DF6/2020\|EPI_ISL_661628\|20 | ···················· | ···················· | ····················· | ···················· | ···················· | ···················· |
| T | 536 hCoV-19/England/QEUH-B67A4A/2020\|EPI_ISL_661629\|20 | ···················· | ···················· | ····················· | ···················· | ···················· | ···················· |
| T | 537 hCoV-19/England/QEUH-B692B9/2020\|EPI_ISL_661630\|20 | ···················· | ···················· | ····················· | ···················· | ···················· | ···················· |
| T | 538 hCoV-19/England/QEUH-B6985D/2020\|EPI_ISL_661631\|20 | ···················· | ···················· | ····················· | ···················· | ···················· | ···················· |
| T | 539 hCoV-19/England/QEUH-B82830/2020\|EPI_ISL_661633\|20 | ···················· | ···················· | ····················· | ···················· | ···················· | ···················· |
| T | 540 hCoV-19/England/QEUH-B82223/2020\|EPI_ISL_661634\|20 | ···················· | ···················· | ····················· | ···················· | ···················· | ···················· |
| T | 541 hCoV-19/England/QEUH-B7EC68/2020\|EPI_ISL_661635\|20 | ···················· | ···················· | ····················· | ···················· | ···················· | ···················· |
| T | 542 hCoV-19/England/QEUH-B82636/2020\|EPI_ISL_661637\|20 | ···················· | ···················· | ····················· | ···················· | ···················· | ···················· |
| T | 543 hCoV-19/England/CAMC-B78527/2020\|EPI_ISL_661640\|20 | ···················· | ···················· | ····················· | ···················· | ···················· | ···················· |
| T | 544 hCoV-19/England/QEUH-B7FE70/2020\|EPI_ISL_661642\|20 | ···················· | ···················· | ····················· | ···················· | ···················· | ···················· |
| T | 545 hCoV-19/England/QEUH-B7F07A/2020\|EPI_ISL_661643\|20 | ···················· | ···················· | ····················· | ···················· | ···················· | ···················· |
| T | 546 hCoV-19/England/QEUH-B8251B/2020\|EPI_ISL_661644\|20 | ···················· | ···················· | ····················· | ···················· | ···················· | ···················· |
| T | 547 hCoV-19/England/QEUH-B7ECB3/2020\|EPI_ISL_661645\|20 | ···················· | ·A·················· | ····················· | ···················· | ···················· | ··A················· |
| T | 548 hCoV-19/England/CAMC-B779C2/2020\|EPI_ISL_661647\|20 | ···················· | ···················· | ····················· | ···················· | ···················· | ···················· |
| T | 549 hCoV-19/England/QEUH-B8052C/2020\|EPI_ISL_661648\|20 | ···················· | ···················· | ····················· | ···················· | ···················· | ···················· |
| T | 550 hCoV-19/England/QEUH-B822F6/2020\|EPI_ISL_661649\|20 | ···················· | ···················· | ····················· | ···················· | ···················· | ···················· |
| T | 551 hCoV-19/England/QEUH-B80A5A/2020\|EPI_ISL_661650\|20 | ···················· | ···················· | ····················· | ···················· | ···················· | ···················· |
| T | 552 hCoV-19/England/CAMC-B783B4/2020\|EPI_ISL_661651\|20 | ···················· | ···················· | ····················· | ···················· | ···················· | ···················· |
| T | 553 hCoV-19/England/CAMC-B782F3/2020\|EPI_ISL_661653\|20 | ···················· | ···················· | ····················· | ···················· | ···················· | ···················· |
| T | 554 hCoV-19/England/QEUH-B82724/2020\|EPI_ISL_661655\|20 | ···················· | ···················· | ····················· | ···················· | ···················· | ···················· |
| T | 555 hCoV-19/England/QEUH-B7F3BD/2020\|EPI_ISL_661658\|20 | ···················· | ···················· | ····················· | ···················· | ···················· | ···················· |
| T | 556 hCoV-19/England/QEUH-B8227E/2020\|EPI_ISL_661659\|20 | ···················· | ···················· | ····················· | ···················· | ···················· | ···················· |
| T | 557 hCoV-19/England/QEUH-B7FF04/2020\|EPI_ISL_661661\|20 | ···················· | ···················· | ····················· | ···················· | ···················· | ···················· |
| T | 558 hCoV-19/England/QEUH-B7ECFF/2020\|EPI_ISL_661663\|20 | ···················· | ···················· | ····················· | ···················· | ···················· | ···················· |
| T | 559 hCoV-19/England/CAMC-B7793B/2020\|EPI_ISL_661664\|20 | ···················· | ···················· | ····················· | ···················· | ···················· | ···················· |
| T | 560 hCoV-19/England/QEUH-B8285E/2020\|EPI_ISL_661665\|20 | ···················· | ···················· | ····················· | ···················· | ···················· | ···················· |
| T | 561 hCoV-19/England/QEUH-B7F3DB/2020\|EPI_ISL_661666\|20 | ···················· | ···················· | ····················· | ···················· | ···················· | ···················· |
| T | 562 hCoV-19/England/QEUH-B81109/2020\|EPI_ISL_661668\|20 | ···················· | ···················· | ····················· | ···················· | ···················· | ···················· |
| T | 563 hCoV-19/England/QEUH-B822C9/2020\|EPI_ISL_661669\|20 | ···················· | ···················· | ····················· | ···················· | ···················· | ···················· |
| T | 564 hCoV-19/England/QEUH-B80A1E/2020\|EPI_ISL_661670\|20 | ···················· | ·A·················· | ····················· | ···················· | ···················· | ···················· |
| T | 565 hCoV-19/England/QEUH-B7FEAD/2020\|EPI_ISL_661671\|20 | ···················· | ···················· | ····················· | ···················· | ···················· | ···················· |
| T | 566 hCoV-19/England/QEUH-B82618/2020\|EPI_ISL_661673\|20 | ···················· | ···················· | ····················· | ···················· | ···················· | ···················· |
| T | 567 hCoV-19/England/QEUH-B807F9/2020\|EPI_ISL_661674\|20 | ···················· | ···················· | ····················· | ···················· | ···················· | ···················· |
| T | 568 hCoV-19/England/CAMC-B778E3/2020\|EPI_ISL_661675\|20 | ···················· | ···················· | ····················· | ···················· | ···················· | ···················· |
| T | 569 hCoV-19/England/QEUH-B7EBF2/2020\|EPI_ISL_661677\|20 | ···················· | ···················· | ····················· | ···················· | ···················· | ···················· |
| T | 570 hCoV-19/England/QEUH-B8236C/2020\|EPI_ISL_661678\|20 | ···················· | ···················· | ····················· | ···················· | ···················· | ···················· |
| T | 571 hCoV-19/England/CAMC-B7835A/2020\|EPI_ISL_661679\|20 | ···················· | ·A·················· | ····················· | ···················· | ···················· | ··A················· |
| T | 572 hCoV-19/England/CAMC-B783A5/2020\|EPI_ISL_661680\|20 | ···················· | ···················· | ····················· | ···················· | ···················· | ···················· |
| T | 573 hCoV-19/England/QEUH-B810DF/2020\|EPI_ISL_661681\|20 | ···················· | ···················· | ····················· | ···················· | ···················· | ···················· |
| T | 574 hCoV-19/England/QEUH-B822D8/2020\|EPI_ISL_661683\|20 | ···················· | ···················· | ····················· | ···················· | ···················· | ···················· |
| T | 575 hCoV-19/England/CAMC-B779FF/2020\|EPI_ISL_661684\|20 | ···················· | ···················· | ····················· | ···················· | ···················· | ···················· |
| T | 576 hCoV-19/England/QEUH-AF49DD/2020\|EPI_ISL_662930\|20 | ···················· | ···················· | ····················· | ···················· | ···················· | ···················· |
| T | 577 hCoV-19/England/QEUH-AF4A34/2020\|EPI_ISL_662931\|20 | ···················· | ···················· | ····················· | ···················· | ···················· | ···················· |
| T | 578 hCoV-19/England/QEUH-AF52E0/2020\|EPI_ISL_662932\|20 | ···················· | ···················· | ····················· | ···················· | ···················· | ···················· |
| T | 579 hCoV-19/England/QEUH-AF3D96/2020\|EPI_ISL_662934\|20 | ···················· | ···················· | ····················· | ···················· | ···················· | ···················· |
| T | 580 hCoV-19/England/QEUH-AF5E0A/2020\|EPI_ISL_662935\|20 | ···················· | ···················· | ····················· | ···················· | ···················· | ···················· |
| T | 581 hCoV-19/England/QEUH-AF49A0/2020\|EPI_ISL_662936\|20 | ···················· | ···················· | ····················· | ···················· | ···················· | ···················· |
| T | 582 hCoV-19/England/QEUH-AF57B4/2020\|EPI_ISL_662937\|20 | ···················· | ···················· | ····················· | ···················· | ······T············· | ···················· |
| T | 583 hCoV-19/England/QEUH-AF5884/2020\|EPI_ISL_662938\|20 | ···················· | ···················· | ····················· | ···················· | ···················· | ···················· |
| T | 584 hCoV-19/England/QEUH-AF4876/2020\|EPI_ISL_662940\|20 | ···················· | ·A·················· | ····················· | ···················· | ···················· | ··A················· |
| T | 585 hCoV-19/England/QEUH-AF3E57/2020\|EPI_ISL_662941\|20 | ···················· | ···················· | ····················· | ···················· | ···················· | ···················· |
| T | 586 hCoV-19/England/QEUH-AF5392/2020\|EPI_ISL_662942\|20 | ···················· | ···················· | ····················· | ···················· | ···················· | ···················· |
| T | 587 hCoV-19/England/QEUH-AF620D/2020\|EPI_ISL_662943\|20 | ···················· | ···················· | ····················· | ···················· | ···················· | ···················· |
| T | 588 hCoV-19/England/QEUH-AF54DB/2020\|EPI_ISL_662944\|20 | ···················· | ···················· | ····················· | ···················· | ···················· | ···················· |
| T | 589 hCoV-19/England/QEUH-AF3E75/2020\|EPI_ISL_662945\|20 | ···················· | ···················· | ····················· | ···················· | ···················· | ···················· |
| T | 590 hCoV-19/England/QEUH-AF6777/2020\|EPI_ISL_662946\|20 | ···················· | ···················· | ····················· | ···················· | ···················· | ···················· |
| T | 591 hCoV-19/England/QEUH-AF5E55/2020\|EPI_ISL_662947\|20 | ···················· | ···················· | ····················· | ···················· | ···················· | ···················· |
| T | 592 hCoV-19/England/QEUH-AF3D4B/2020\|EPI_ISL_662948\|20 | ···················· | ···················· | ····················· | ···················· | ···················· | ···················· |
| T | 593 hCoV-19/England/QEUH-AF4DD1/2020\|EPI_ISL_662949\|20 | ···················· | ···················· | ····················· | ···················· | ···················· | ···················· |
| T | 594 hCoV-19/England/QEUH-AF4CB6/2020\|EPI_ISL_662950\|20 | ···················· | ···················· | ····················· | ···················· | ···················· | ···················· |
| T | 595 hCoV-19/England/QEUH-AF5013/2020\|EPI_ISL_662951\|20 | ···················· | ···················· | ····················· | ···················· | ···················· | ···················· |
| T | 596 hCoV-19/England/QEUH-AF53A1/2020\|EPI_ISL_662955\|20 | ···················· | ···················· | ····················· | ···················· | ···················· | ···················· |

| | | | | | | | |
|---|---|---|---|---|---|---|---|
| T | 597 hCoV-19/England/QEUH-AF5769/2020\|EPI_ISL_662956\|20 | ···················· | ···················· | ····················· | ···················· | ···················· | ···················· |
| T | 598 hCoV-19/England/QEUH-AF5AAC/2020\|EPI_ISL_662957\|20 | ···················· | ···················· | ····················· | ···················· | ···················· | ···················· |
| T | 599 hCoV-19/England/QEUH-AF667A/2020\|EPI_ISL_662960\|20 | ···················· | ···················· | ····················· | ···················· | ···················· | ···················· |
| T | 600 hCoV-19/England/QEUH-AF4CD4/2020\|EPI_ISL_662961\|20 | ···················· | ···················· | ····················· | ···················· | ···················· | ···················· |
| T | 601 hCoV-19/England/QEUH-AF514D/2020\|EPI_ISL_662963\|20 | ···················· | ···················· | ····················· | ···················· | ···················· | ···················· |
| T | 602 hCoV-19/England/QEUH-AF520E/2020\|EPI_ISL_662964\|20 | ···················· | ···················· | ····················· | ···················· | ···················· | ···················· |
| T | 603 hCoV-19/England/QEUH-AF4E65/2020\|EPI_ISL_662965\|20 | ···················· | ···················· | ····················· | ···················· | ···················· | ···················· |
| T | 604 hCoV-19/England/QEUH-AF53CF/2020\|EPI_ISL_662966\|20 | ···················· | ···················· | ····················· | ···················· | ···················· | ···················· |
| T | 605 hCoV-19/England/QEUH-AF56E4/2020\|EPI_ISL_662967\|20 | ···················· | ···················· | ····················· | ···················· | ···················· | ···················· |
| T | 606 hCoV-19/England/QEUH-AF509B/2020\|EPI_ISL_662968\|20 | ···················· | ···················· | ····················· | ···················· | ···················· | ···················· |
| T | 607 hCoV-19/England/QEUH-AF3E93/2020\|EPI_ISL_662969\|20 | ···················· | ···················· | ····················· | ···················· | ···················· | ···················· |
| T | 608 hCoV-19/England/QEUH-AF672C/2020\|EPI_ISL_662970\|20 | ···················· | ···················· | ····················· | ···················· | ···················· | ···················· |
| T | 609 hCoV-19/England/QEUH-AF665C/2020\|EPI_ISL_662971\|20 | ···················· | ···················· | ····················· | ···················· | ···················· | ···················· |
| T | 610 hCoV-19/England/QEUH-AF68FC/2020\|EPI_ISL_662972\|20 | ···················· | ···················· | ····················· | ···················· | ···················· | ···················· |
| T | 611 hCoV-19/England/QEUH-AF4BF5/2020\|EPI_ISL_662973\|20 | ···················· | ···················· | ····················· | ···················· | ···················· | ···················· |
| T | 612 hCoV-19/England/QEUH-AF3DA5/2020\|EPI_ISL_662974\|20 | ···················· | ···················· | ····················· | ···················· | ···················· | ···················· |
| T | 613 hCoV-19/England/QEUH-AF69CC/2020\|EPI_ISL_662975\|20 | ···················· | ···················· | ····················· | ···················· | ···················· | ···················· |
| T | 614 hCoV-19/England/QEUH-AF52FF/2020\|EPI_ISL_662976\|20 | ···················· | ···················· | ····················· | ···················· | ···················· | ···················· |
| T | 615 hCoV-19/England/QEUH-AF69AE/2020\|EPI_ISL_662977\|20 | ···················· | ···················· | ····················· | ···················· | ···················· | ···················· |
| T | 616 hCoV-19/England/QEUH-AF55AB/2020\|EPI_ISL_662978\|20 | ···················· | ···················· | ····················· | ···················· | ···················· | ···················· |
| T | 617 hCoV-19/England/QEUH-AF621C/2020\|EPI_ISL_662980\|20 | ···················· | ···················· | ····················· | ···················· | ······T············· | ···················· |
| T | 618 hCoV-19/England/QEUH-AF4894/2020\|EPI_ISL_662981\|20 | ···················· | ···················· | ····················· | ···················· | ···················· | ···················· |
| T | 619 hCoV-19/England/QEUH-AF5B21/2020\|EPI_ISL_662982\|20 | ···················· | ···················· | ····················· | ···················· | ···················· | ···················· |
| T | 620 hCoV-19/England/QEUH-AF4D86/2020\|EPI_ISL_662983\|20 | ···················· | ···················· | ····················· | ···················· | ···················· | ···················· |
| T | 621 hCoV-19/England/QEUH-AF5040/2020\|EPI_ISL_662986\|20 | ···················· | ···················· | ····················· | ···················· | ···················· | ···················· |
| T | 622 hCoV-19/England/QEUH-AF3D2D/2020\|EPI_ISL_662987\|20 | ···················· | ···················· | ····················· | ···················· | ···················· | ···················· |
| T | 623 hCoV-19/England/QEUH-AF55BA/2020\|EPI_ISL_662988\|20 | ···················· | ···················· | ····················· | ···················· | ···················· | ···················· |
| T | 624 hCoV-19/England/QEUH-AF568A/2020\|EPI_ISL_662989\|20 | ···················· | ···················· | ····················· | ···················· | ···················· | ···················· |
| T | 625 hCoV-19/England/QEUH-AF56A8/2020\|EPI_ISL_662990\|20 | ···················· | ·A·················· | ····················· | ···················· | ···················· | ···A················ |
| T | 626 hCoV-19/England/QEUH-AF5D85/2020\|EPI_ISL_662991\|20 | ···················· | ···················· | ····················· | ···················· | ···················· | ···················· |
| T | 627 hCoV-19/England/QEUH-AF51E3/2020\|EPI_ISL_662992\|20 | ···················· | ·A·················· | ····················· | ···················· | ···················· | ···················· |
| T | 628 hCoV-19/England/QEUH-AF572D/2020\|EPI_ISL_662993\|20 | ···················· | ···················· | ····················· | ···················· | ···················· | ···················· |
| T | 629 hCoV-19/England/QEUH-AF6786/2020\|EPI_ISL_662994\|20 | ···················· | ···················· | ····················· | ···················· | ···················· | ···················· |
| T | 630 hCoV-19/England/QEUH-AF664D/2020\|EPI_ISL_662995\|20 | ···················· | ···················· | ····················· | ···················· | ···················· | ···················· |
| T | 631 hCoV-19/England/QEUH-AF522C/2020\|EPI_ISL_662996\|20 | ···················· | ···················· | ····················· | ···················· | ···················· | ···················· |
| T | 632 hCoV-19/England/QEUH-AF4DA4/2020\|EPI_ISL_662997\|20 | ···················· | ···················· | ····················· | ···················· | ···················· | ···················· |
| T | 633 hCoV-19/England/QEUH-AF409C/2020\|EPI_ISL_662998\|20 | ···················· | ···················· | ····················· | ···················· | ······T············· | ···················· |
| T | 634 hCoV-19/England/QEUH-AF674A/2020\|EPI_ISL_662999\|20 | ···················· | ···················· | ····················· | ···················· | ···················· | ···················· |
| T | 635 hCoV-19/England/LOND-1250BB0/2020\|EPI_ISL_665252\|2 | ···················· | ···················· | ····················· | ···················· | ······T············· | ···················· |
| T | 637 hCoV-19/Norway/CS-05/2020\|EPI_ISL_665254\|2020-04-1 | ···················· | ···················· | ····················· | ···················· | ···················· | ···················· |
| T | 638 hCoV-19/Norway/CS-06/2020\|EPI_ISL_665255\|2020-04-2 | ···················· | ···················· | ····················· | ···················· | ···················· | ···················· |
| T | 639 hCoV-19/Wales/PHWC-48D122/2020\|EPI_ISL_666583\|2020 | ···················· | ···················· | ····················· | ···················· | ···················· | ···················· |
| T | 640 hCoV-19/Wales/PHWC-48D131/2020\|EPI_ISL_666584\|2020 | ···················· | ···················· | ····················· | ···················· | ···················· | ···················· |
| T | 641 hCoV-19/Wales/PHWC-48D140/2020\|EPI_ISL_666585\|2020 | ···················· | ···················· | ····················· | ···················· | ···················· | ···················· |
| T | 642 hCoV-19/Wales/PHWC-48D15F/2020\|EPI_ISL_666586\|2020 | ···················· | ···················· | ····················· | ···················· | ···················· | ···················· |
| T | 643 hCoV-19/Wales/PHWC-48D16E/2020\|EPI_ISL_666587\|2020 | ···················· | ···················· | ····················· | ···················· | ···················· | ···················· |
| T | 644 hCoV-19/Wales/PHWC-48D17D/2020\|EPI_ISL_666588\|2020 | ···················· | ···················· | ····················· | ···················· | ···················· | ···················· |
| T | 645 hCoV-19/Wales/PHWC-48D19B/2020\|EPI_ISL_666589\|2020 | ···················· | ···················· | ····················· | ···················· | ···················· | ···················· |
| T | 646 hCoV-19/Australia/NSW1200/2020\|EPI_ISL_667800\|2020 | ···················· | ···················· | ····················· | ···················· | ···················· | ···················· |
| T | 647 hCoV-19/Australia/NSW1201/2020\|EPI_ISL_667801\|2020 | ···················· | ·A·················· | ···A················· | ···················· | ···················· | ···················· |
| T | 648 hCoV-19/Australia/NSW1203/2020\|EPI_ISL_667802\|2020 | ···················· | ···················· | ····················· | ···················· | ···················· | ···················· |
| T | 649 hCoV-19/Australia/NSW1205/2020\|EPI_ISL_667803\|2020 | ···················· | ···················· | ····················· | ···················· | ···················· | ···················· |
| T | 650 hCoV-19/Australia/NSW1206/2020\|EPI_ISL_667804\|2020 | ···················· | ···················· | ····················· | ···················· | ···················· | ···················· |
| T | 651 hCoV-19/Australia/NSW1207/2020\|EPI_ISL_667805\|2020 | ···················· | ·A·················· | ····················· | ···················· | ···················· | ···A················ |
| T | 652 hCoV-19/Australia/NSW1208/2020\|EPI_ISL_667806\|2020 | ···················· | ···················· | ····················· | ···················· | ···················· | ···················· |
| T | 653 hCoV-19/Taiwan/CGMH-CGU-34/2020\|EPI_ISL_667807\|202 | ···················· | ···················· | ····················· | ···················· | ···················· | ···················· |
| T | 654 hCoV-19/Taiwan/CGMH-CGU-35/2020\|EPI_ISL_667808\|202 | ···················· | ···················· | ····················· | ···················· | ···················· | ···················· |
| T | 655 hCoV-19/Taiwan/CGMH-CGU-36/2020\|EPI_ISL_667809\|202 | ···················· | ·A·················· | ····················· | ···················· | ···················· | ···A················ |
| T | 656 hCoV-19/England/CAMC-B79517/2020\|EPI_ISL_667812\|20 | ···················· | ···················· | ····················· | ···················· | ···················· | ···················· |
| T | 657 hCoV-19/England/CAMC-B79D1F/2020\|EPI_ISL_667813\|20 | ···················· | ···················· | ····················· | ···················· | ···················· | ···················· |
| T | 658 hCoV-19/England/QEUH-B7EB4D/2020\|EPI_ISL_667814\|20 | ···················· | ·A·················· | ····················· | ···················· | ···················· | ···A················ |
| T | 659 hCoV-19/England/CAMC-B78ADD/2020\|EPI_ISL_667815\|20 | ···················· | ···················· | ····················· | ···················· | ···················· | ···················· |
| T | 660 hCoV-19/England/QEUH-B7ED92/2020\|EPI_ISL_667816\|20 | ···················· | ·A·················· | ····················· | ···················· | ···················· | ···················· |
| T | 661 hCoV-19/England/CAMC-B79B24/2020\|EPI_ISL_667817\|20 | ···················· | ···················· | ····················· | ···················· | ······T············· | ···················· |
| T | 662 hCoV-19/England/CAMC-B796BA/2020\|EPI_ISL_667819\|20 | ···················· | ···················· | ····················· | ···················· | ···················· | ···················· |
| T | 663 hCoV-19/England/CAMC-B789C1/2020\|EPI_ISL_667820\|20 | ···················· | ···················· | ····················· | ···················· | ···················· | ···················· |
| T | 664 hCoV-19/England/CAMC-B7A9DE/2020\|EPI_ISL_667821\|20 | ···················· | ·A·················· | ····················· | ···················· | ···················· | ···················· |
| T | 665 hCoV-19/Denmark/DCGC-11570/2020\|EPI_ISL_671000\|202 | ···················· | ···················· | ····················· | ···················· | ···················· | ···················· |
| T | 666 hCoV-19/Denmark/DCGC-11571/2020\|EPI_ISL_671001\|202 | ···················· | ···················· | ····················· | ···················· | ···················· | ···················· |
| T | 667 hCoV-19/Denmark/DCGC-11631/2020\|EPI_ISL_671037\|202 | ···················· | ···················· | ····················· | ···················· | ···················· | ···················· |
| T | 668 hCoV-19/Denmark/DCGC-11633/2020\|EPI_ISL_671039\|202 | ···················· | ···················· | ····················· | ···················· | ···················· | ···················· |
| T | 669 hCoV-19/Denmark/DCGC-11647/2020\|EPI_ISL_671046\|202 | ···················· | ·A·················· | ····················· | ···················· | ···················· | ···················· |
| T | 670 hCoV-19/Denmark/DCGC-11648/2020\|EPI_ISL_671047\|202 | ···················· | ···················· | ····················· | ···················· | ···················· | ···················· |
| T | 671 hCoV-19/Denmark/DCGC-11650/2020\|EPI_ISL_671048\|202 | ···················· | ···················· | ····················· | ···················· | ···················· | ···················· |
| T | 672 hCoV-19/Denmark/DCGC-11651/2020\|EPI_ISL_671049\|202 | ···················· | ···················· | ····················· | ···················· | ···················· | ···················· |
| T | 673 hCoV-19/Denmark/DCGC-11655/2020\|EPI_ISL_671050\|202 | ···················· | ···················· | ····················· | ···················· | ···················· | ···················· |
| T | 674 hCoV-19/Denmark/DCGC-11657/2020\|EPI_ISL_671051\|202 | ···················· | ···················· | ····················· | ···················· | ···················· | ···················· |
| T | 675 hCoV-19/Denmark/DCGC-11658/2020\|EPI_ISL_671052\|202 | ···················· | ·A·················· | ····················· | ···················· | ···················· | ···················· |
| T | 676 hCoV-19/Denmark/DCGC-11660/2020\|EPI_ISL_671053\|202 | ···················· | ···················· | ····················· | ···················· | ···················· | ···················· |
| T | 677 hCoV-19/Denmark/DCGC-11661/2020\|EPI_ISL_671054\|202 | ···················· | ···················· | ····················· | ···················· | ···················· | ···················· |

```
T 678 hCoV-19/Denmark/DCGC-11662/2020|EPI_ISL_671055|202 ···················· ···················· ···················· ···················· ···················· ····················
T 679 hCoV-19/Denmark/DCGC-11667/2020|EPI_ISL_671056|202 ···················· ···················· ···················· ···················· ···················· ····················
T 680 hCoV-19/Denmark/DCGC-11669/2020|EPI_ISL_671057|202 ···················· ···················· ···················· ···················· ···················· ····················
T 681 hCoV-19/Denmark/DCGC-11670/2020|EPI_ISL_671058|202 ···················· ···················· ···················· ···················· ···················· ····················
T 682 hCoV-19/Denmark/DCGC-11673/2020|EPI_ISL_671059|202 ···················· ···················· ···················· ···················· ···················· ····················
T 683 hCoV-19/Denmark/DCGC-11674/2020|EPI_ISL_671060|202 ···················· ·A·················· ···················· ···················· ···················· ····················
T 684 hCoV-19/Denmark/DCGC-11675/2020|EPI_ISL_671061|202 ···················· ···················· ···················· ···················· ···················· ····················
T 685 hCoV-19/Denmark/DCGC-11679/2020|EPI_ISL_671062|202 ···················· ···················· ···················· ···················· ···················· ····················
T 686 hCoV-19/Denmark/DCGC-11680/2020|EPI_ISL_671063|202 ···················· ···················· ···················· ···················· ···················· ····················
T 687 hCoV-19/Denmark/DCGC-11681/2020|EPI_ISL_671064|202 ···················· ···················· ···················· ···················· ···················· ····················
T 688 hCoV-19/Denmark/DCGC-11682/2020|EPI_ISL_671065|202 ···················· ···················· ···················· ···················· ···················· ····················
T 689 hCoV-19/Denmark/DCGC-11683/2020|EPI_ISL_671066|202 ···················· ···················· ···················· ···················· ···················· ····················
T 690 hCoV-19/Denmark/DCGC-11684/2020|EPI_ISL_671067|202 ···················· ···················· ···················· ···················· ···················· ····················
T 691 hCoV-19/Denmark/DCGC-11687/2020|EPI_ISL_671068|202 ···················· ···················· ···················· ···················· ···················· ····················
T 692 hCoV-19/Denmark/DCGC-11688/2020|EPI_ISL_671069|202 ···················· ···················· ···················· ···················· ···················· ····················
T 693 hCoV-19/Denmark/DCGC-11689/2020|EPI_ISL_671070|202 ···················· ···················· ···················· ···················· ···················· ····················
T 694 hCoV-19/Denmark/DCGC-11690/2020|EPI_ISL_671071|202 ···················· ···················· ···················· ···················· ···················· ····················
T 695 hCoV-19/Denmark/DCGC-11691/2020|EPI_ISL_671072|202 ···················· ···················· ···················· ···················· ···················· ····················
T 696 hCoV-19/Denmark/DCGC-11692/2020|EPI_ISL_671073|202 ···················· ···················· ···················· ···················· ···················· ····················
T 697 hCoV-19/Denmark/DCGC-11693/2020|EPI_ISL_671074|202 ···················· ···················· ···················· ···················· ···················· ····················
T 698 hCoV-19/Denmark/DCGC-11694/2020|EPI_ISL_671075|202 ···················· ···················· ···················· ···················· ···················· ····················
T 699 hCoV-19/Denmark/DCGC-11697/2020|EPI_ISL_671076|202 ···················· ···················· ···················· ···················· ···················· ····················
T 700 hCoV-19/Denmark/DCGC-11698/2020|EPI_ISL_671077|202 ···················· ···················· ···················· ···················· ···················· ····················
T 701 hCoV-19/Denmark/DCGC-11699/2020|EPI_ISL_671078|202 ···················· ···················· ···················· ···················· ···················· ····················
T 702 hCoV-19/Denmark/DCGC-11701/2020|EPI_ISL_671079|202 ···················· ···················· ···················· ···················· ···················· ····················
T 703 hCoV-19/Denmark/DCGC-11702/2020|EPI_ISL_671080|202 ···················· ···················· ···················· ···················· ···················· ····················
T 704 hCoV-19/Denmark/DCGC-11703/2020|EPI_ISL_671081|202 ···················· ·A·················· ···················· ···················· ···················· ····················
T 705 hCoV-19/Denmark/DCGC-11704/2020|EPI_ISL_671082|202 ···················· ···················· ···················· ···················· ···················· ····················
T 706 hCoV-19/Denmark/DCGC-11706/2020|EPI_ISL_671083|202 ···················· ···················· ···················· ···················· ···················· ····················
T 707 hCoV-19/Denmark/DCGC-11707/2020|EPI_ISL_671084|202 ···················· ···················· ···················· ···················· ···················· ····················
T 708 hCoV-19/Denmark/DCGC-11708/2020|EPI_ISL_671085|202 ···················· ···················· ···················· ···················· ···················· ····················
T 709 hCoV-19/Denmark/DCGC-11709/2020|EPI_ISL_671086|202 ···················· ···················· ···················· ···················· ···················· ····················
T 710 hCoV-19/Denmark/DCGC-11711/2020|EPI_ISL_671087|202 ···················· ···················· ···················· ···················· ···················· ····················
T 711 hCoV-19/Denmark/DCGC-11713/2020|EPI_ISL_671088|202 ···················· ···················· ···················· ···················· ·····T·············· ····················
T 712 hCoV-19/Denmark/DCGC-11714/2020|EPI_ISL_671089|202 ···················· ···················· ···················· ···················· ···················· ····················
T 713 hCoV-19/USA/CA-CZB-13014/2020|EPI_ISL_672300|2020- ···················· ·A·················· ···················· ···················· ···················· ····················
T 714 hCoV-19/USA/CA-CZB-13016/2020|EPI_ISL_672301|2020- ···················· ·A·················· ···················· ···················· ···················· ····················
T 715 hCoV-19/USA/CA-CZB-13017/2020|EPI_ISL_672302|2020- ···················· ·A·················· ···················· ···················· ·NNN················ ···················· 1
T 717 hCoV-19/USA/CA-CZB-13019/2020|EPI_ISL_672304|2020- ···················· ·A·················· ···················· ···················· ···················· ····················
T 720 hCoV-19/USA/CA-CZB-13023/2020|EPI_ISL_672307|2020- ···················· ···················· ···················· ···················· ···················· ····················
T 721 hCoV-19/USA/CA-CZB-13025/2020|EPI_ISL_672308|2020- ···················· ···················· ···················· ···················· ···················· ····················
T 722 hCoV-19/USA/CA-CZB-13026/2020|EPI_ISL_672309|2020- ···················· ···················· ···················· ···················· ···················· ····················
T 723 hCoV-19/USA/CA-CZB-13027/2020|EPI_ISL_672310|2020- ···················· ···················· ···················· ···················· ···················· ····················
T 724 hCoV-19/USA/CA-CZB-13028/2020|EPI_ISL_672311|2020- ···················· ···················· ···················· ···················· ···················· ····················
T 726 hCoV-19/USA/CA-CZB-13030/2020|EPI_ISL_672313|2020- ···················· ···················· ···················· ···················· ···················· ····················
T 728 hCoV-19/USA/CA-CZB-13035/2020|EPI_ISL_672315|2020- ···················· ···················· ···················· ···················· ···················· ····················
T 729 hCoV-19/USA/CA-CZB-13036/2020|EPI_ISL_672316|2020- ···················· ···················· ···················· ···················· ···················· ····················
T 731 hCoV-19/USA/CA-CZB-13038/2020|EPI_ISL_672318|2020- ···················· ···················· ···················· ···················· ···················· ····················
T 732 hCoV-19/USA/CA-CZB-13039/2020|EPI_ISL_672319|2020- ···················· ···················· ···················· ···················· ···················· ····················
T 733 hCoV-19/USA/CA-CZB-13043/2020|EPI_ISL_672320|2020- ···················· ···················· ···················· ···················· ···················· ····················
T 734 hCoV-19/USA/CA-CZB-13044/2020|EPI_ISL_672321|2020- ···················· ···················· ···················· ···················· ···················· ····················
T 735 hCoV-19/USA/CA-CZB-13045/2020|EPI_ISL_672322|2020- ···················· ···················· ···················· ···················· ···················· ····················
T 736 hCoV-19/USA/CA-CZB-13046/2020|EPI_ISL_672323|2020- ···················· ···················· ···················· ···················· ···················· ····················
T 737 hCoV-19/USA/CA-CZB-13047/2020|EPI_ISL_672324|2020- ···················· ···················· ···················· ···················· ···················· ····················
T 738 hCoV-19/USA/CA-CZB-13048/2020|EPI_ISL_672325|2020- ···················· ···················· ···················· ···················· ···················· ····················
T 739 hCoV-19/USA/CA-CZB-13052/2020|EPI_ISL_672326|2020- ···················· ···················· ···················· ···················· ···················· ····················
T 740 hCoV-19/USA/CA-CZB-13054/2020|EPI_ISL_672327|2020- ···················· ···················· ···················· ···················· ···················· ····················
T 742 hCoV-19/USA/CA-CZB-13059/2020|EPI_ISL_672329|2020- ···················· ···················· ···················· ···················· ···················· ····················
T 743 hCoV-19/USA/CA-CZB-13060/2020|EPI_ISL_672330|2020- ···················· ···················· ···················· ···················· ···················· ····················
T 746 hCoV-19/USA/CA-CZB-12960/2020|EPI_ISL_672377|2020- ···················· ···················· ···················· ···················· ···················· ····················
T 747 hCoV-19/USA/CA-CZB-12961/2020|EPI_ISL_672378|2020- ···················· ···················· ···················· ···················· ···················· ····················
T 748 hCoV-19/USA/CA-CZB-12964/2020|EPI_ISL_672379|2020- ···················· ···················· ···················· ···················· ···················· ····················
T 749 hCoV-19/USA/CA-CZB-12965/2020|EPI_ISL_672380|2020- ···················· ···················· ···················· ···················· ···················· ····················
T 750 hCoV-19/USA/CA-CZB-12966/2020|EPI_ISL_672381|2020- ···················· ···················· ···················· ···················· ···················· ····················
T 751 hCoV-19/USA/CA-CZB-12967/2020|EPI_ISL_672382|2020- ···················· ···················· ···················· ···················· ···················· ····················
T 752 hCoV-19/USA/CA-CZB-12968/2020|EPI_ISL_672383|2020- ···················· ···················· ···················· ···················· ···················· ····················
T 753 hCoV-19/USA/CA-CZB-12970/2020|EPI_ISL_672384|2020- ···················· ···················· ···················· ···················· ···················· ····················
T 754 hCoV-19/USA/CA-CZB-12971/2020|EPI_ISL_672385|2020- ···················· ···················· ···················· ···················· N··················· ····················
T 755 hCoV-19/USA/CA-CZB-5845/2020|EPI_ISL_672386|2020-0 ···················· ···················· ···················· ···················· ···················· ····················
T 756 hCoV-19/USA/CA-CZB-5841/2020|EPI_ISL_672387|2020-0 ···················· ···················· ···················· ···················· ···················· ····················
T 757 hCoV-19/USA/CA-CZB-5710/2020|EPI_ISL_672388|2020-0 ···················· ·A·················· ···················· ···················· ···················· ···········N········
T 760 hCoV-19/USA/CA-CZB-5687/2020|EPI_ISL_672391|2020-0 ···················· ···················· ···················· ···················· ···················· ····················
T 762 hCoV-19/USA/CA-CZB-5160/2020|EPI_ISL_672393|2020-0 ···················· ·A·················· ···················· ···················· ···················· ····················
T 763 hCoV-19/USA/CA-CZB-13143/2020|EPI_ISL_672394|2020- ···················· ·A·················· ···················· ···················· ···················· ····················
T 764 hCoV-19/USA/CA-CZB-13142/2020|EPI_ISL_672395|2020- ···················· ·A·················· ···················· ···················· ···················· ····················
T 765 hCoV-19/USA/CA-CZB-13141/2020|EPI_ISL_672396|2020- ···················· ···················· ···················· ···················· ···················· ····················
T 766 hCoV-19/USA/CA-CZB-13140/2020|EPI_ISL_672397|2020- ···················· ···················· ···················· ···················· ···················· ····················
T 767 hCoV-19/USA/CA-CZB-13139/2020|EPI_ISL_672398|2020- ···················· ·A·················· ···················· ···················· ···················· ····················
T 768 hCoV-19/USA/CA-CZB-13138/2020|EPI_ISL_672399|2020- ···················· ·A·················· ···················· ···················· ···················· ····················
T 769 hCoV-19/England/QEUH-B8139A/2020|EPI_ISL_661685|20 ···················· ···················· ···················· ···················· ···················· ····················
```

| | | | | | | | | |
|---|---|---|---|---|---|---|---|---|
| T | 770 | hCoV-19/England/QEUH-B82557/2020\|EPI_ISL_661686\|20 | ···················· | ···················· | ····················· | ···················· | ···················· | ···················· |
| T | 771 | hCoV-19/England/QEUH-B80489/2020\|EPI_ISL_661688\|20 | ···················· | ···················· | ····················· | ···················· | ···················· | ···················· |
| T | 772 | hCoV-19/England/QEUH-B810C0/2020\|EPI_ISL_661689\|20 | ···················· | ···················· | ····················· | ···················· | ···················· | ···················· |
| T | 773 | hCoV-19/England/QEUH-B8135E/2020\|EPI_ISL_661690\|20 | ···················· | ···················· | ····················· | ···················· | ···················· | ···················· |
| T | 774 | hCoV-19/England/CAMC-B7791D/2020\|EPI_ISL_661691\|20 | ···················· | ···················· | ····················· | ···················· | ······T············· | ···················· |
| T | 775 | hCoV-19/England/CAMC-B7830F/2020\|EPI_ISL_661693\|20 | ···················· | ···················· | ····················· | ···················· | ···················· | ···················· |
| T | 776 | hCoV-19/England/QEUH-B82575/2020\|EPI_ISL_661694\|20 | ···················· | ···················· | ····················· | ···················· | ···················· | ···················· |
| T | 777 | hCoV-19/England/CAMC-B78572/2020\|EPI_ISL_661695\|20 | ···················· | ···················· | ····················· | ···················· | ···················· | ···················· |
| T | 778 | hCoV-19/England/CAMC-B785AF/2020\|EPI_ISL_661696\|20 | ···················· | ···················· | ····················· | ···················· | ······T············· | ···················· |
| T | 779 | hCoV-19/England/CAMC-B78387/2020\|EPI_ISL_661697\|20 | ···················· | ···················· | ····················· | ···················· | ···················· | ···················· |
| T | 780 | hCoV-19/England/QEUH-B7EAD7/2020\|EPI_ISL_661698\|20 | ···················· | ···················· | ····················· | ···················· | ···················· | ···················· |
| T | 781 | hCoV-19/England/QEUH-B827BB/2020\|EPI_ISL_661699\|20 | ···················· | ···················· | ····················· | ···················· | ···················· | ···················· |
| T | 782 | hCoV-19/England/QEUH-AF5ECE/2020\|EPI_ISL_662920\|20 | ···················· | ···················· | ····················· | ···················· | ···················· | ···················· |
| T | 783 | hCoV-19/England/EXET-13AEC6/2020\|EPI_ISL_665100\|20 | ···················· | ···················· | ····················· | ···················· | ······T············· | ···················· |
| T | 784 | hCoV-19/England/EXET-13AED5/2020\|EPI_ISL_665101\|20 | ···················· | ···················· | ····················· | ···················· | ······T············· | ···················· |
| T | 785 | hCoV-19/England/EXET-13AEF3/2020\|EPI_ISL_665102\|20 | ···················· | ···················· | ····················· | ···················· | ······T············· | ···················· |
| T | 786 | hCoV-19/England/EXET-13AF3C/2020\|EPI_ISL_665103\|20 | ···················· | ···················· | ····················· | ···················· | ······T············· | ···················· |
| T | 787 | hCoV-19/England/EXET-13AFA5/2020\|EPI_ISL_665104\|20 | ···················· | ···················· | ····················· | ···················· | ······T············· | ···················· |
| T | 788 | hCoV-19/England/EXET-13AFB4/2020\|EPI_ISL_665105\|20 | ···················· | ···················· | ····················· | ···················· | ······T············· | ···················· |
| T | 789 | hCoV-19/England/EXET-13B01A/2020\|EPI_ISL_665106\|20 | ···················· | ···················· | ····················· | ···················· | ···················· | ···················· |
| T | 790 | hCoV-19/England/EXET-13B038/2020\|EPI_ISL_665107\|20 | ···················· | ···················· | ····················· | ···················· | ···················· | ···················· |
| T | 791 | hCoV-19/England/EXET-13B092/2020\|EPI_ISL_665108\|20 | ···················· | ···················· | ····················· | ···················· | ······T············· | ···················· |
| T | 792 | hCoV-19/England/EXET-13B0A1/2020\|EPI_ISL_665109\|20 | ···················· | ···················· | ····················· | ···················· | ······T············· | ···················· |
| T | 793 | hCoV-19/England/EXET-13B0B0/2020\|EPI_ISL_665110\|20 | ···················· | ···················· | ····················· | ···················· | ······T············· | ···················· |
| T | 794 | hCoV-19/England/EXET-13B126/2020\|EPI_ISL_665111\|20 | ···················· | ···················· | ····················· | ···················· | ······T············· | ···················· |
| T | 795 | hCoV-19/England/EXET-13B180/2020\|EPI_ISL_665112\|20 | ···················· | ···················· | ····················· | ···················· | ······T············· | ···················· |
| T | 796 | hCoV-19/England/LIVE-DB991D/2020\|EPI_ISL_665113\|20 | ···················· | ···················· | ····················· | ···················· | ···················· | ···················· |
| T | 797 | hCoV-19/England/LIVE-DBAC22/2020\|EPI_ISL_665114\|20 | ···················· | ···················· | ····················· | ···················· | ···················· | ···················· |
| T | 798 | hCoV-19/England/LIVE-DBACF5/2020\|EPI_ISL_665115\|20 | ···················· | ···················· | ····················· | ···················· | ···················· | ···················· |
| T | 799 | hCoV-19/England/LIVE-DBAD01/2020\|EPI_ISL_665116\|20 | ···················· | ···················· | ····················· | ···················· | ···················· | ···················· |
| T | 800 | hCoV-19/England/LIVE-DBAD10/2020\|EPI_ISL_665117\|20 | ···················· | ···················· | ····················· | ···················· | ···················· | ···················· |
| T | 801 | hCoV-19/England/LIVE-DBAD98/2020\|EPI_ISL_665118\|20 | ···················· | ···················· | ····················· | ···················· | ···················· | ···················· |
| T | 802 | hCoV-19/England/LIVE-DBADA7/2020\|EPI_ISL_665119\|20 | ···················· | ···················· | ····················· | ···················· | ···················· | ···················· |
| T | 803 | hCoV-19/England/LIVE-DBADB6/2020\|EPI_ISL_665120\|20 | ···················· | ···················· | ····················· | ···················· | ···················· | ···················· |
| T | 804 | hCoV-19/England/LIVE-DBAEA4/2020\|EPI_ISL_665121\|20 | ···················· | ···················· | ····················· | ···················· | ···················· | ···················· |
| T | 805 | hCoV-19/England/LIVE-DBAED1/2020\|EPI_ISL_665122\|20 | ···················· | ···················· | ····················· | ···················· | ···················· | ···················· |
| T | 806 | hCoV-19/England/LIVE-DBB016/2020\|EPI_ISL_665123\|20 | ···················· | ···················· | ····················· | ···················· | ···················· | ···················· |
| T | 807 | hCoV-19/England/LIVE-DBB59F/2020\|EPI_ISL_665124\|20 | ···················· | ···················· | ····················· | ···················· | ···················· | ···················· |
| T | 808 | hCoV-19/England/LIVE-DBB67E/2020\|EPI_ISL_665125\|20 | ···················· | ···················· | ····················· | ···················· | ···A················ | ···················· |
| T | 809 | hCoV-19/England/LIVE-DBB6BA/2020\|EPI_ISL_665126\|20 | ···················· | ···················· | ····················· | ···················· | ···················· | ···················· |
| T | 810 | hCoV-19/England/LIVE-DBB6F6/2020\|EPI_ISL_665127\|20 | ···················· | ···················· | ····················· | ···················· | ···················· | ···················· |
| T | 811 | hCoV-19/Wales/PHWC-48C18D/2020\|EPI_ISL_666400\|2020 | ···················· | ···················· | ····················· | ···················· | ···················· | ···················· |
| T | 812 | hCoV-19/Wales/PHWC-48C19C/2020\|EPI_ISL_666401\|2020 | ···················· | ···················· | ····················· | ···················· | ···················· | ···················· |
| T | 813 | hCoV-19/Wales/PHWC-48C1AB/2020\|EPI_ISL_666402\|2020 | ···················· | ···················· | ····················· | ···················· | ···················· | ···················· |
| T | 814 | hCoV-19/Wales/PHWC-48C1BA/2020\|EPI_ISL_666403\|2020 | ···················· | ···················· | ····················· | ···················· | ···················· | ···················· |
| T | 815 | hCoV-19/Wales/PHWC-48C1C9/2020\|EPI_ISL_666404\|2020 | ···················· | ···················· | ····················· | ···················· | ···················· | ···················· |
| T | 816 | hCoV-19/Wales/PHWC-48C1E7/2020\|EPI_ISL_666405\|2020 | ···················· | ···················· | ····················· | ···················· | ···················· | ···················· |
| T | 817 | hCoV-19/Wales/PHWC-48C1F6/2020\|EPI_ISL_666406\|2020 | ···················· | ···················· | ····················· | ···················· | ···················· | ···················· |
| T | 818 | hCoV-19/Wales/PHWC-48C211/2020\|EPI_ISL_666407\|2020 | ···················· | ···················· | ····················· | ···················· | ···················· | ···················· |
| T | 819 | hCoV-19/Wales/PHWC-48C220/2020\|EPI_ISL_666408\|2020 | ···················· | ···················· | ····················· | ···················· | ···················· | ···················· |
| T | 820 | hCoV-19/Wales/PHWC-48C23F/2020\|EPI_ISL_666409\|2020 | ···················· | ···················· | ····················· | ···················· | ···················· | ···················· |
| T | 821 | hCoV-19/Wales/PHWC-48C25D/2020\|EPI_ISL_666410\|2020 | ···················· | ···················· | ····················· | ···················· | ···A················ | ···················· |
| T | 822 | hCoV-19/Wales/PHWC-48C27B/2020\|EPI_ISL_666411\|2020 | ···················· | ···················· | ····················· | ···················· | ···················· | ···················· |
| T | 823 | hCoV-19/Wales/PHWC-48C299/2020\|EPI_ISL_666412\|2020 | ···················· | ···················· | ····················· | ···················· | ···················· | ···················· |
| T | 824 | hCoV-19/Wales/PHWC-48C2B7/2020\|EPI_ISL_666413\|2020 | ···················· | ···················· | ····················· | ···················· | ···A················ | ···················· |
| T | 825 | hCoV-19/Wales/PHWC-48C2C6/2020\|EPI_ISL_666414\|2020 | ···················· | ···················· | ····················· | ···················· | ···················· | ···················· |
| T | 826 | hCoV-19/Wales/PHWC-48C2E4/2020\|EPI_ISL_666415\|2020 | ···················· | ···················· | ····················· | ···················· | ···················· | ···················· |
| T | 827 | hCoV-19/Wales/PHWC-48C2F3/2020\|EPI_ISL_666416\|2020 | ···················· | ···················· | ····················· | ···················· | ···················· | ···················· |
| T | 828 | hCoV-19/Wales/PHWC-48C30F/2020\|EPI_ISL_666417\|2020 | ···················· | ···················· | ····················· | ···················· | ···················· | ···················· |
| T | 829 | hCoV-19/Wales/PHWC-48C31E/2020\|EPI_ISL_666418\|2020 | ···················· | ···················· | ····················· | ···················· | ···················· | ···················· |
| T | 830 | hCoV-19/Wales/PHWC-48C32D/2020\|EPI_ISL_666419\|2020 | ···················· | ···················· | ····················· | ···················· | ···················· | ···················· |
| T | 831 | hCoV-19/Wales/PHWC-48C34B/2020\|EPI_ISL_666420\|2020 | ···················· | ···················· | ····················· | ···················· | ···················· | ···················· |
| T | 832 | hCoV-19/Wales/PHWC-48C369/2020\|EPI_ISL_666421\|2020 | ···················· | ···················· | ····················· | ···················· | ···················· | ···················· |
| T | 833 | hCoV-19/Wales/PHWC-48C378/2020\|EPI_ISL_666422\|2020 | ···················· | ···················· | ····················· | ···················· | ···················· | ···················· |
| T | 834 | hCoV-19/Wales/PHWC-48C3B4/2020\|EPI_ISL_666423\|2020 | ···················· | ···················· | ····················· | ···················· | ···················· | ···················· |
| T | 835 | hCoV-19/Wales/PHWC-48C3C3/2020\|EPI_ISL_666424\|2020 | ···················· | ···················· | ····················· | ···················· | ···················· | ···················· |
| T | 836 | hCoV-19/Wales/PHWC-48C3D2/2020\|EPI_ISL_666425\|2020 | ···················· | ···················· | ····················· | ···················· | ···················· | ···················· |
| T | 837 | hCoV-19/Wales/PHWC-48C3E1/2020\|EPI_ISL_666426\|2020 | ···················· | ···················· | ····················· | ···················· | ···················· | ···················· |
| T | 838 | hCoV-19/Wales/PHWC-48C3F0/2020\|EPI_ISL_666427\|2020 | ···················· | ···················· | ····················· | ···················· | ···················· | ···················· |
| T | 839 | hCoV-19/Wales/PHWC-48C40C/2020\|EPI_ISL_666428\|2020 | ···················· | ···················· | ····················· | ···················· | ···················· | ···················· |
| T | 840 | hCoV-19/Wales/PHWC-48C448/2020\|EPI_ISL_666429\|2020 | ···················· | ···················· | ····················· | ···················· | ···················· | ···················· |
| T | 843 | hCoV-19/Wales/PHWC-48C493/2020\|EPI_ISL_666432\|2020 | ···················· | ···················· | ····················· | ···················· | ···················· | ···················· |
| T | 844 | hCoV-19/Wales/PHWC-48C4A2/2020\|EPI_ISL_666433\|2020 | ···················· | ···················· | ····················· | ···················· | ···················· | ···················· |
| T | 845 | hCoV-19/Wales/PHWC-48C4C0/2020\|EPI_ISL_666434\|2020 | ···················· | ···················· | ····················· | ···················· | ···················· | ···················· |
| T | 846 | hCoV-19/Wales/PHWC-48C4DF/2020\|EPI_ISL_666435\|2020 | ···················· | ···················· | ····················· | ···················· | ···················· | ···················· |
| T | 847 | hCoV-19/Wales/PHWC-48C4EE/2020\|EPI_ISL_666436\|2020 | ···················· | ···················· | ····················· | ···················· | ···················· | ···················· |
| T | 848 | hCoV-19/Wales/PHWC-48C509/2020\|EPI_ISL_666437\|2020 | ···················· | ···················· | ····················· | ···················· | ···················· | ···················· |
| T | 849 | hCoV-19/Wales/PHWC-48C527/2020\|EPI_ISL_666438\|2020 | ···················· | ···················· | ····················· | ···················· | ···················· | ···················· |
| T | 850 | hCoV-19/Wales/PHWC-48C536/2020\|EPI_ISL_666439\|2020 | ···················· | ···················· | ····················· | ···················· | ···················· | ···················· |
| T | 851 | hCoV-19/Wales/PHWC-48C554/2020\|EPI_ISL_666440\|2020 | ···················· | ···················· | ····················· | ···················· | ···················· | ···················· |

| | | | | | | | |
|---|---|---|---|---|---|---|---|
| T | 853 hCoV-19/Wales/PHWC-48C572/2020\|EPI_ISL_666442\|2020 | .................... | .................... | ..................... | .................... | .................... | .................... |
| T | 854 hCoV-19/Wales/PHWC-48C581/2020\|EPI_ISL_666443\|2020 | .................... | .................... | ..................... | .................... | .................... | .................... |
| T | 855 hCoV-19/Wales/PHWC-48C590/2020\|EPI_ISL_666444\|2020 | .................... | .................... | ..................... | .................... | .................... | .................... |
| T | 856 hCoV-19/Wales/PHWC-48C5BE/2020\|EPI_ISL_666445\|2020 | .................... | .................... | ..................... | .................... | .................... | .................... |
| T | 857 hCoV-19/Wales/PHWC-48C5CD/2020\|EPI_ISL_666446\|2020 | .................... | .................... | ..................... | .................... | .................... | .................... |
| T | 858 hCoV-19/Wales/PHWC-48C5EB/2020\|EPI_ISL_666447\|2020 | .................... | .................... | ..................... | .................... | .................... | .................... |
| T | 859 hCoV-19/Wales/PHWC-48C606/2020\|EPI_ISL_666448\|2020 | .................... | .................... | ..................... | .................... | .................... | .................... |
| T | 860 hCoV-19/Wales/PHWC-48C615/2020\|EPI_ISL_666449\|2020 | .................... | .................... | ..................... | .................... | .................... | .................... |
| T | 861 hCoV-19/Wales/PHWC-48C633/2020\|EPI_ISL_666450\|2020 | .................... | .................... | ..................... | .................... | .................... | .................... |
| T | 862 hCoV-19/Wales/PHWC-48C651/2020\|EPI_ISL_666451\|2020 | .................... | .................... | ..................... | .................... | .................... | .................... |
| T | 864 hCoV-19/Wales/PHWC-48C67F/2020\|EPI_ISL_666453\|2020 | .................... | .................... | ..................... | .................... | .................... | .................... |
| T | 865 hCoV-19/Wales/PHWC-48C68E/2020\|EPI_ISL_666454\|2020 | .................... | .................... | ..................... | .................... | .................... | .................... |
| T | 866 hCoV-19/Wales/PHWC-48C69D/2020\|EPI_ISL_666455\|2020 | .................... | .................... | ..................... | .................... | .................... | .................... |
| T | 867 hCoV-19/Wales/PHWC-48C6AC/2020\|EPI_ISL_666456\|2020 | .................... | .................... | ..................... | .................... | .................... | .................... |
| T | 868 hCoV-19/Wales/PHWC-48C6BB/2020\|EPI_ISL_666457\|2020 | .................... | .................... | ..................... | .................... | .................... | .................... |
| T | 869 hCoV-19/Wales/PHWC-48C6E8/2020\|EPI_ISL_666458\|2020 | .................... | .................... | ..................... | .................... | .................... | .................... |
| T | 870 hCoV-19/Japan/IC-0273/2020\|EPI_ISL_667702\|2020-11 | .................... | ............C....... | ..................... | .................... | .................... | .................... |
| T | 871 hCoV-19/Japan/IC-0274/2020\|EPI_ISL_667703\|2020-11 | .................... | ............C....... | ..................... | .................... | .................... | .................... |
| T | 872 hCoV-19/Japan/IC-0275/2020\|EPI_ISL_667704\|2020-11 | .................... | ............C....... | ..................... | .................... | .................... | .................... |
| T | 873 hCoV-19/Japan/IC-0276/2020\|EPI_ISL_667705\|2020-11 | .................... | .A.................. | ..................... | .................... | .................... | ...A................ |
| T | 874 hCoV-19/Japan/IC-0277/2020\|EPI_ISL_667706\|2020-11 | .................... | ............C....... | ..................... | .................... | .................... | .................... |
| T | 875 hCoV-19/Japan/IC-0278/2020\|EPI_ISL_667707\|2020-11 | .................... | .A.................. | ..................... | .................... | .................... | ...A................ |
| T | 876 hCoV-19/Japan/IC-0279/2020\|EPI_ISL_667708\|2020-11 | .................... | .................... | ..................... | .................... | .................... | .................... |
| T | 877 hCoV-19/Japan/IC-0280/2020\|EPI_ISL_667709\|2020-11 | .................... | .................... | ..................... | .................... | .................... | .................... |
| T | 878 hCoV-19/Japan/IC-0281/2020\|EPI_ISL_667710\|2020-11 | .................... | ............C....... | ..................... | .................... | .................... | .................... |
| T | 879 hCoV-19/Japan/IC-0282/2020\|EPI_ISL_667711\|2020-11 | .................... | ............C....... | ..................... | .................... | .................... | .................... |
| T | 880 hCoV-19/Japan/IC-0283/2020\|EPI_ISL_667712\|2020-11 | .................... | .................... | ..................... | .................... | .................... | .................... |
| T | 881 hCoV-19/Japan/IC-0284/2020\|EPI_ISL_667713\|2020-11 | .................... | .................... | ..................... | .................... | .................... | .................... |
| T | 882 hCoV-19/Japan/IC-0285/2020\|EPI_ISL_667714\|2020-11 | .................... | ............C....... | ..................... | .................... | .................... | .................... |
| T | 883 hCoV-19/Japan/IC-0286/2020\|EPI_ISL_667715\|2020-11 | .................... | ............C....... | ..................... | .................... | .................... | .................... |
| T | 884 hCoV-19/Japan/IC-0287/2020\|EPI_ISL_667716\|2020-11 | .................... | .................... | ..................... | .................... | .................... | .................... |
| T | 885 hCoV-19/Japan/IC-0288/2020\|EPI_ISL_667717\|2020-11 | .................... | ....A............... | ..................... | .................... | .................... | .................... |
| T | 886 hCoV-19/Japan/IC-0289/2020\|EPI_ISL_667718\|2020-11 | .................... | .................... | ..................... | .................... | .................... | .................... |
| T | 887 hCoV-19/Japan/IC-0290/2020\|EPI_ISL_667719\|2020-11 | .................... | .................... | ..................... | .................... | .................... | .................... |
| T | 888 hCoV-19/Japan/IC-0291/2020\|EPI_ISL_667720\|2020-11 | .................... | .................... | ..................... | .................... | .................... | .................... |
| T | 889 hCoV-19/Japan/IC-0292/2020\|EPI_ISL_667721\|2020-11 | .................... | .................... | ..................... | .................... | .................... | .................... |
| T | 890 hCoV-19/Japan/IC-0293/2020\|EPI_ISL_667722\|2020-11 | .................... | .A.................. | ..................... | .................... | .................... | ...A................ |
| T | 891 hCoV-19/Japan/IC-0294/2020\|EPI_ISL_667723\|2020-11 | .................... | ....A............... | ..................... | .................... | .................... | .................... |
| T | 892 hCoV-19/Japan/IC-0295/2020\|EPI_ISL_667724\|2020-11 | .................... | ......A............. | ..................... | .................... | .................... | .................... |
| T | 893 hCoV-19/Japan/IC-0296/2020\|EPI_ISL_667725\|2020-11 | .................... | .................... | ..................... | .................... | .................... | .................... |
| T | 894 hCoV-19/Japan/IC-0297/2020\|EPI_ISL_667726\|2020-11 | .................... | .................... | ..................... | .................... | .................... | .................... |
| T | 895 hCoV-19/Japan/IC-0298/2020\|EPI_ISL_667727\|2020-11 | .................... | .................... | ..................... | .................... | .................... | .................... |
| T | 896 hCoV-19/Japan/IC-0299/2020\|EPI_ISL_667728\|2020-11 | .................... | .................... | ..................... | .................... | .................... | .................... |
| T | 897 hCoV-19/Japan/IC-0301/2020\|EPI_ISL_667730\|2020-11 | .................... | .................... | ..................... | .................... | .................... | .................... |
| T | 898 hCoV-19/Japan/IC-0302/2020\|EPI_ISL_667731\|2020-11 | .................... | .A.................. | ..................... | .................... | .................... | .................... |
| T | 899 hCoV-19/Japan/IC-0303/2020\|EPI_ISL_667732\|2020-11 | .................... | .................... | ..................... | .................... | .................... | .................... |
| T | 900 hCoV-19/Japan/IC-0304/2020\|EPI_ISL_667733\|2020-11 | .................... | .................... | ..................... | .................... | .................... | .................... |
| T | 901 hCoV-19/Japan/IC-0306/2020\|EPI_ISL_667735\|2020-09 | .................... | .................... | ..................... | .................... | .................... | .................... |
| T | 902 hCoV-19/Japan/IC-0307/2020\|EPI_ISL_667736\|2020-09 | .................... | .................... | ..................... | .................... | .................... | .................... |
| T | 903 hCoV-19/Japan/IC-0308/2020\|EPI_ISL_667737\|2020-10 | .................... | .A.......A.......... | ..................... | .................... | .................... | ...A................ |
| T | 904 hCoV-19/Japan/IC-0309/2020\|EPI_ISL_667738\|2020-10 | .................... | .................... | ..................... | .................... | .................... | .................... |
| T | 905 hCoV-19/Japan/IC-0310/2020\|EPI_ISL_667739\|2020-10 | .................... | .................... | ..................... | .................... | .................... | .................... |
| T | 906 hCoV-19/Japan/IC-0311/2020\|EPI_ISL_667740\|2020-11 | .................... | .A.................. | ..................... | .................... | .................... | ...A................ |
| T | 907 hCoV-19/Japan/IC-0312/2020\|EPI_ISL_667741\|2020-11 | .................... | .................... | ..................... | .................... | .................... | .................... |
| T | 908 hCoV-19/Japan/IC-0313/2020\|EPI_ISL_667742\|2020-11 | .................... | .................... | ..................... | .................... | .................... | .................... |
| T | 909 hCoV-19/Japan/IC-0314/2020\|EPI_ISL_667743\|2020-11 | .................... | .................... | ..................... | .................... | .................... | .................... |
| T | 910 hCoV-19/Japan/IC-0315/2020\|EPI_ISL_667744\|2020-11 | .................... | .................... | ..................... | .................... | .................... | .................... |
| T | 911 hCoV-19/Japan/IC-0316/2020\|EPI_ISL_667745\|2020-11 | .................... | .................... | ..................... | .................... | .................... | .................... |
| T | 912 hCoV-19/Japan/IC-0317/2020\|EPI_ISL_667746\|2020-11 | .................... | .................... | ..................... | .................... | .................... | .................... |
| T | 913 hCoV-19/Japan/IC-0318/2020\|EPI_ISL_667747\|2020-11 | .................... | .................... | ..................... | .................... | .................... | .................... |
| T | 914 hCoV-19/Japan/IC-0319/2020\|EPI_ISL_667748\|2020-11 | .................... | .................... | ..................... | .................... | .................... | .................... |
| T | 915 hCoV-19/Japan/IC-0320/2020\|EPI_ISL_667749\|2020-11 | .................... | .................... | ..................... | .................... | .................... | .................... |
| T | 916 hCoV-19/Japan/IC-0321/2020\|EPI_ISL_667750\|2020-11 | .................... | .................... | ..................... | .................... | .................... | .................... |
| T | 917 hCoV-19/Japan/IC-0322/2020\|EPI_ISL_667751\|2020-11 | .................... | .................... | ..................... | .................... | .................... | .................... |
| T | 918 hCoV-19/Japan/IC-0323/2020\|EPI_ISL_667752\|2020-11 | .................... | .................... | ..................... | .................... | .................... | .................... |
| T | 919 hCoV-19/Japan/IC-0324/2020\|EPI_ISL_667753\|2020-11 | .................... | .................... | ..................... | .................... | .................... | .................... |
| T | 920 hCoV-19/Japan/IC-0325/2020\|EPI_ISL_667754\|2020-11 | .................... | .................... | ..................... | .................... | .................... | .................... |
| T | 921 hCoV-19/Japan/IC-0326/2020\|EPI_ISL_667755\|2020-08 | .................... | .................... | ..................... | .................... | .................... | .................... |
| T | 922 hCoV-19/Japan/IC-0327/2020\|EPI_ISL_667756\|2020-08 | .................... | .................... | ..................... | .................... | .................... | .................... |
| T | 923 hCoV-19/Japan/IC-0328/2020\|EPI_ISL_667757\|2020-09 | .................... | .................... | ..................... | .................... | .................... | .................... |
| T | 924 hCoV-19/Canada/UN-CRCHUM-PreFreeze-2/2020\|EPI_ISL_ | .................... | .................... | ..................... | .................... | .................... | .................... |
| T | 925 hCoV-19/Canada/UN-CRCHUM-PreFreeze-5/2020\|EPI_ISL_ | .................... | .................... | ..................... | .................... | .................... | .................... |
| T | 926 hCoV-19/Canada/UN-CRCHUM-PostFreeze-5/2020\|EPI_ISL | .................... | .................... | ..................... | .................... | .................... | .................... |
| T | 927 hCoV-19/Australia/NSW1172/2020\|EPI_ISL_667777\|2020 | .................... | .................... | ..................... | .................... | .................... | .................... |
| T | 928 hCoV-19/Australia/NSW1173/2020\|EPI_ISL_667778\|2020 | .................... | .................... | ..................... | .................... | .................... | .................... |
| T | 929 hCoV-19/Australia/NSW1174/2020\|EPI_ISL_667779\|2020 | .................... | .................... | ..................... | .................... | .................... | .................... |
| T | 930 hCoV-19/Australia/NSW1176/2020\|EPI_ISL_667780\|2020 | .................... | .................... | ..................... | .................... | .................... | .................... |
| T | 931 hCoV-19/Australia/NSW1177/2020\|EPI_ISL_667781\|2020 | .................... | .................... | ..................... | .................... | .................... | .................... |
| T | 932 hCoV-19/Australia/NSW1178/2020\|EPI_ISL_667782\|2020 | .................... | .................... | ..................... | .................... | .................... | .................... |
| T | 933 hCoV-19/Australia/NSW1180/2020\|EPI_ISL_667783\|2020 | .................... | .................... | ..................... | .................... | .................... | .................... |

| | | | | | | | | |
|---|---|---|---|---|---|---|---|---|
| T | 934 | hCoV-19/Australia/NSW1181/2020\|EPI_ISL_667784\|2020 | ···················· | ···················· | ····················· | ···················· | ···················· | ···················· |
| T | 935 | hCoV-19/Australia/NSW1182/2020\|EPI_ISL_667785\|2020 | ···················· | ·A·················· | ····················· | ···················· | ···················· | ··A················· |
| T | 936 | hCoV-19/Australia/NSW1184/2020\|EPI_ISL_667786\|2020 | ···················· | ···················· | ····················· | ···················· | ···················· | ···················· |
| T | 937 | hCoV-19/Australia/NSW1186/2020\|EPI_ISL_667787\|2020 | ···················· | ···················· | ····················· | ···················· | ···················· | ···················· |
| T | 938 | hCoV-19/Australia/NSW1187/2020\|EPI_ISL_667788\|2020 | ···················· | ···················· | ····················· | ···················· | ···················· | ···················· |
| T | 939 | hCoV-19/Denmark/DCGC-11576/2020\|EPI_ISL_671002\|202 | ···················· | ···················· | ····················· | ···················· | ···················· | ···················· |
| T | 940 | hCoV-19/Denmark/DCGC-11577/2020\|EPI_ISL_671003\|202 | ···················· | ···················· | ····················· | ···················· | ···················· | ···················· |
| T | 941 | hCoV-19/Denmark/DCGC-11580/2020\|EPI_ISL_671004\|202 | ···················· | ···················· | ····················· | ···················· | ···················· | ···················· |
| T | 942 | hCoV-19/Denmark/DCGC-11582/2020\|EPI_ISL_671005\|202 | ···················· | ···················· | ····················· | ···················· | ···················· | ···················· |
| T | 943 | hCoV-19/Denmark/DCGC-11583/2020\|EPI_ISL_671006\|202 | ···················· | ···················· | ····················· | ···················· | ···················· | ···················· |
| T | 944 | hCoV-19/Denmark/DCGC-11584/2020\|EPI_ISL_671007\|202 | ···················· | ···················· | ····················· | ···················· | ···················· | ···················· |
| T | 945 | hCoV-19/Denmark/DCGC-11589/2020\|EPI_ISL_671008\|202 | ···················· | ···················· | ····················· | ···················· | ···················· | ···················· |
| T | 946 | hCoV-19/Denmark/DCGC-11590/2020\|EPI_ISL_671009\|202 | ···················· | ···················· | ····················· | ···················· | ···················· | ···················· |
| T | 947 | hCoV-19/Denmark/DCGC-11591/2020\|EPI_ISL_671010\|202 | ···················· | ·A·················· | ····················· | ···················· | ···················· | ··A················· |
| T | 948 | hCoV-19/Denmark/DCGC-11592/2020\|EPI_ISL_671011\|202 | ···················· | ···················· | ····················· | ···················· | ···················· | ···················· |
| T | 949 | hCoV-19/Denmark/DCGC-11593/2020\|EPI_ISL_671012\|202 | ···················· | ···················· | ····················· | ···················· | ···················· | ···················· |
| T | 950 | hCoV-19/Denmark/DCGC-11594/2020\|EPI_ISL_671013\|202 | ···················· | ···················· | ····················· | ···················· | ···················· | ···················· |
| T | 951 | hCoV-19/Denmark/DCGC-11596/2020\|EPI_ISL_671014\|202 | ···················· | ···················· | ····················· | ···················· | ···················· | ···················· |
| T | 952 | hCoV-19/Denmark/DCGC-11598/2020\|EPI_ISL_671015\|202 | ···················· | ···················· | ····················· | ···················· | ···················· | ···················· |
| T | 953 | hCoV-19/Denmark/DCGC-11601/2020\|EPI_ISL_671016\|202 | ···················· | ···················· | ····················· | ···················· | ···················· | ···················· |
| T | 954 | hCoV-19/Denmark/DCGC-11602/2020\|EPI_ISL_671017\|202 | ···················· | ···················· | ····················· | ···················· | ···················· | ···················· |
| T | 955 | hCoV-19/Denmark/DCGC-11603/2020\|EPI_ISL_671018\|202 | ···················· | ···················· | ····················· | ···················· | ···················· | ···················· |
| T | 956 | hCoV-19/Denmark/DCGC-11604/2020\|EPI_ISL_671019\|202 | ···················· | ···················· | ····················· | ···················· | ···················· | ···················· |
| T | 957 | hCoV-19/Denmark/DCGC-11606/2020\|EPI_ISL_671020\|202 | ···················· | ···················· | ····················· | ···················· | ···················· | ···················· |
| T | 958 | hCoV-19/Denmark/DCGC-11607/2020\|EPI_ISL_671021\|202 | ···················· | ···················· | ····················· | ···················· | ···················· | ···················· |
| T | 959 | hCoV-19/Denmark/DCGC-11609/2020\|EPI_ISL_671022\|202 | ···················· | ···················· | ····················· | ···················· | ···················· | ···················· |
| T | 960 | hCoV-19/Denmark/DCGC-11613/2020\|EPI_ISL_671023\|202 | ···················· | ···················· | ····················· | ···················· | ···················· | ···················· |
| T | 961 | hCoV-19/Denmark/DCGC-11614/2020\|EPI_ISL_671024\|202 | ···················· | ···················· | ····················· | ···················· | ···················· | ···················· |
| T | 962 | hCoV-19/Denmark/DCGC-11615/2020\|EPI_ISL_671025\|202 | ···················· | ···················· | ····················· | ···················· | ···················· | ···················· |
| T | 963 | hCoV-19/Denmark/DCGC-11616/2020\|EPI_ISL_671026\|202 | ···················· | ···················· | ····················· | ···················· | ···················· | ···················· |
| T | 964 | hCoV-19/Denmark/DCGC-11618/2020\|EPI_ISL_671027\|202 | ···················· | ···················· | ····················· | ···················· | ···················· | ···················· |
| T | 965 | hCoV-19/Denmark/DCGC-11619/2020\|EPI_ISL_671028\|202 | ···················· | ···················· | ····················· | ···················· | ···················· | ···················· |
| T | 966 | hCoV-19/Denmark/DCGC-11620/2020\|EPI_ISL_671029\|202 | ···················· | ···················· | ····················· | ···················· | ···················· | ···················· |
| T | 967 | hCoV-19/Denmark/DCGC-11621/2020\|EPI_ISL_671030\|202 | ···················· | ·····C·············· | ····················· | ···················· | ···················· | ···················· |
| T | 968 | hCoV-19/Denmark/DCGC-11622/2020\|EPI_ISL_671031\|202 | ···················· | ···················· | ····················· | ···················· | ···················· | ···················· |
| T | 969 | hCoV-19/Denmark/DCGC-11623/2020\|EPI_ISL_671032\|202 | ···················· | ···················· | ····················· | ···················· | ···················· | ···················· |
| T | 970 | hCoV-19/Denmark/DCGC-11625/2020\|EPI_ISL_671033\|202 | ···················· | ···················· | ····················· | ···················· | ···················· | ···················· |
| T | 971 | hCoV-19/Denmark/DCGC-11626/2020\|EPI_ISL_671034\|202 | ···················· | ···················· | ····················· | ···················· | ···················· | ···················· |
| T | 972 | hCoV-19/Denmark/DCGC-11627/2020\|EPI_ISL_671035\|202 | ···················· | ···················· | ····················· | ···················· | ···················· | ···················· |
| T | 973 | hCoV-19/Denmark/DCGC-11629/2020\|EPI_ISL_671036\|202 | ···················· | ·A·················· | ····················· | ···················· | ···················· | ··A················· |
| T | 974 | hCoV-19/Denmark/DCGC-11632/2020\|EPI_ISL_671038\|202 | ···················· | ···················· | ····················· | ···················· | ···················· | ···················· |
| T | 975 | hCoV-19/Denmark/DCGC-11636/2020\|EPI_ISL_671040\|202 | ···················· | ···················· | ····················· | ···················· | ···················· | ···················· |
| T | 976 | hCoV-19/Denmark/DCGC-11639/2020\|EPI_ISL_671041\|202 | ···················· | ···················· | ····················· | ···················· | ···················· | ···················· |
| T | 977 | hCoV-19/Denmark/DCGC-11640/2020\|EPI_ISL_671042\|202 | ···················· | ···················· | ····················· | ···················· | ···················· | ···················· |
| T | 978 | hCoV-19/Denmark/DCGC-11643/2020\|EPI_ISL_671043\|202 | ···················· | ···················· | ····················· | ···················· | ···················· | ···················· |
| T | 979 | hCoV-19/Denmark/DCGC-11644/2020\|EPI_ISL_671044\|202 | ···················· | ···················· | ····················· | ···················· | ···················· | ···················· |
| T | 980 | hCoV-19/Denmark/DCGC-11645/2020\|EPI_ISL_671045\|202 | ···················· | ···················· | ····················· | ···················· | ···················· | ···················· |
| T | 981 | hCoV-19/USA/CA-CZB-13063/2020\|EPI_ISL_672333\|2020- | ···················· | ···················· | ····················· | ···················· | ···················· | ···················· |
| T | 982 | hCoV-19/USA/CA-CZB-13064/2020\|EPI_ISL_672334\|2020- | ···················· | ···················· | ····················· | ···················· | ···················· | ···················· |
| T | 983 | hCoV-19/USA/CA-CZB-13066/2020\|EPI_ISL_672335\|2020- | ···················· | ···················· | ····················· | ···················· | ···················· | ···················· |
| T | 984 | hCoV-19/USA/CA-CZB-13067/2020\|EPI_ISL_672336\|2020- | ···················· | ···················· | ····················· | ···················· | ···················· | ···················· |
| T | 985 | hCoV-19/USA/CA-CZB-13068/2020\|EPI_ISL_672337\|2020- | ···················· | ···················· | ····················· | ···················· | ···················· | ···················· |
| T | 986 | hCoV-19/USA/CA-CZB-13069/2020\|EPI_ISL_672338\|2020- | ···················· | ···················· | ····················· | ···················· | ···················· | ···················· |
| T | 987 | hCoV-19/USA/CA-CZB-13071/2020\|EPI_ISL_672339\|2020- | ···················· | ···················· | ····················· | ···················· | ···················· | ···················· |
| T | 988 | hCoV-19/USA/CA-CZB-13072/2020\|EPI_ISL_672340\|2020- | ···················· | ···················· | ····················· | ···················· | ···················· | ···················· |
| T | 989 | hCoV-19/USA/CA-CZB-13073/2020\|EPI_ISL_672341\|2020- | ···················· | ···················· | ····················· | ···················· | ···················· | ···················· |
| T | 993 | hCoV-19/USA/CA-CZB-13087/2020\|EPI_ISL_672345\|2020- | ···················· | ···················· | ····················· | ···················· | ···················· | ···················· |
| T | 995 | hCoV-19/USA/CA-CZB-13091/2020\|EPI_ISL_672347\|2020- | ···················· | ···················· | ····················· | ···················· | ···················· | ···················· |
| T | 997 | hCoV-19/USA/CA-CZB-13095/2020\|EPI_ISL_672349\|2020- | ···················· | ···················· | ····················· | ···················· | ···················· | ···················· |
| T | 999 | hCoV-19/USA/CA-CZB-13098/2020\|EPI_ISL_672351\|2020- | ···················· | ···················· | ····················· | ···················· | ···················· | ···················· |
| T | 1000 | hCoV-19/USA/CA-CZB-13099/2020\|EPI_ISL_672352\|2020- | ···················· | ···················· | ····················· | ···················· | ···················· | ···················· |
| T | 1001 | hCoV-19/USA/CA-CZB-13100/2020\|EPI_ISL_672353\|2020- | ···················· | ···················· | ····················· | ···················· | ···················· | ···················· |
| T | 1002 | hCoV-19/USA/CA-CZB-5810/2020\|EPI_ISL_672354\|2020-0 | ···················· | ···················· | ····················· | ···················· | ···················· | ···················· |
| T | 1004 | hCoV-19/USA/CA-CZB-5764/2020\|EPI_ISL_672356\|2020-0 | ···················· | ···················· | ····················· | ···················· | ·M·················· | ···················· |
| T | 1005 | hCoV-19/USA/CA-CZB-5762/2020\|EPI_ISL_672357\|2020-0 | ···················· | ···················· | ····················· | ···················· | ···················· | ···················· |
| T | 1006 | hCoV-19/USA/CA-CZB-5757/2020\|EPI_ISL_672358\|2020-0 | ···················· | ·A·················· | ····················· | ···················· | ···················· | ···················· |
| T | 1007 | hCoV-19/USA/CA-CZB-5751/2020\|EPI_ISL_672359\|2020-0 | ···················· | ···················· | ····················· | ···················· | ···················· | ···················· |
| T | 1008 | hCoV-19/USA/CA-CZB-12931/2020\|EPI_ISL_672360\|2020- | ···················· | ·A·················· | ····················· | ···················· | ···················· | ···················· |
| T | 1009 | hCoV-19/USA/CA-CZB-12933/2020\|EPI_ISL_672361\|2020- | ···················· | ···················· | ····················· | ···················· | ···················· | ···················· |
| T | 1010 | hCoV-19/USA/CA-CZB-12936/2020\|EPI_ISL_672362\|2020- | ···················· | ···················· | ····················· | ···················· | ···················· | ···················· |
| T | 1011 | hCoV-19/USA/CA-CZB-12938/2020\|EPI_ISL_672363\|2020- | ···················· | ···················· | ····················· | ···················· | ···················· | ···················· |
| T | 1012 | hCoV-19/USA/CA-CZB-12940/2020\|EPI_ISL_672364\|2020- | ···················· | ···················· | ····················· | ···················· | ···················· | ···················· |
| T | 1013 | hCoV-19/USA/CA-CZB-12943/2020\|EPI_ISL_672365\|2020- | ···················· | ···················· | ····················· | ···················· | ···················· | ···················· |
| T | 1014 | hCoV-19/USA/CA-CZB-12944/2020\|EPI_ISL_672366\|2020- | ···················· | ·A·················· | ····················· | ···················· | ···················· | ···················· |
| T | 1015 | hCoV-19/USA/CA-CZB-12945/2020\|EPI_ISL_672367\|2020- | ···················· | ···················· | ····················· | ···················· | ···················· | ···················· |
| T | 1016 | hCoV-19/USA/CA-CZB-12946/2020\|EPI_ISL_672368\|2020- | ···················· | ···················· | ····················· | ···················· | ···················· | ···················· |
| T | 1017 | hCoV-19/USA/CA-CZB-12947/2020\|EPI_ISL_672369\|2020- | ···················· | ···················· | ····················· | ···················· | ···················· | ···················· |
| T | 1018 | hCoV-19/USA/CA-CZB-12948/2020\|EPI_ISL_672370\|2020- | ···················· | ···················· | ····················· | ···················· | ···················· | ···················· |
| T | 1019 | hCoV-19/USA/CA-CZB-12950/2020\|EPI_ISL_672371\|2020- | ···················· | ···················· | ····················· | ···················· | ···················· | ···················· |
| T | 1020 | hCoV-19/USA/CA-CZB-12951/2020\|EPI_ISL_672372\|2020- | ···················· | ···················· | ····················· | ···················· | ···················· | ···················· |

| | | | | | | | |
|---|---|---|---|---|---|---|---|
| T | 1021 hCoV-19/USA/CA-CZB-12952/2020\|EPI_ISL_672373\|2020- | ···················· | ···················· | ····················· | ···················· | ···················· | ···················· |
| T | 1022 hCoV-19/USA/CA-CZB-12956/2020\|EPI_ISL_672374\|2020- | ···················· | ···················· | ····················· | ···················· | ···················· | ···················· |
| T | 1023 hCoV-19/USA/CA-CZB-12957/2020\|EPI_ISL_672375\|2020- | ···················· | ···················· | ····················· | ···················· | ···················· | ···················· |
| T | 1024 hCoV-19/USA/CA-CZB-12958/2020\|EPI_ISL_672376\|2020- | ···················· | ···················· | ····················· | ···················· | ···················· | ···················· |
| T | 1025 hCoV-19/England/QEUH-B66674/2020\|EPI_ISL_661500\|20 | ···················· | ···················· | ····················· | ···················· | ···················· | ···················· |
| T | 1026 hCoV-19/England/QEUH-B67A3B/2020\|EPI_ISL_661501\|20 | ···················· | ···················· | ····················· | ···················· | ···················· | ···················· |
| T | 1027 hCoV-19/England/QEUH-B665C2/2020\|EPI_ISL_661502\|20 | ···················· | ···················· | ····················· | ···················· | ···················· | ···················· |
| T | 1028 hCoV-19/England/QEUH-B6934D/2020\|EPI_ISL_661503\|20 | ···················· | ·A·················· | ····················· | ···················· | ···················· | ···················· |
| T | 1029 hCoV-19/England/QEUH-B6C632/2020\|EPI_ISL_661504\|20 | ···················· | ···················· | ····················· | ···················· | ···················· | ···················· |
| T | 1030 hCoV-19/England/QEUH-B662E9/2020\|EPI_ISL_661510\|20 | ···················· | ···················· | ····················· | ···················· | ···················· | ···················· |
| T | 1031 hCoV-19/England/QEUH-B67A86/2020\|EPI_ISL_661512\|20 | ···················· | ···················· | ····················· | ···················· | ···················· | ···················· |
| T | 1032 hCoV-19/England/QEUH-B670A2/2020\|EPI_ISL_661514\|20 | ···················· | ···················· | ····················· | ···················· | ···················· | ···················· |
| T | 1033 hCoV-19/England/QEUH-B6895B/2020\|EPI_ISL_661517\|20 | ···················· | ···················· | ····················· | ···················· | ···················· | ···················· |
| T | 1034 hCoV-19/England/QEUH-B67C26/2020\|EPI_ISL_661518\|20 | ···················· | ···················· | ····················· | ···················· | ···················· | ···················· |
| T | 1035 hCoV-19/England/QEUH-B67A0E/2020\|EPI_ISL_661519\|20 | ···················· | ···················· | ····················· | ···················· | ···················· | ···················· |
| T | 1036 hCoV-19/England/QEUH-B69A57/2020\|EPI_ISL_661521\|20 | ···················· | ···················· | ····················· | ···················· | ···················· | ···················· |
| T | 1037 hCoV-19/England/QEUH-B6795C/2020\|EPI_ISL_661522\|20 | ···················· | ·A·················· | ····················· | ···················· | ···················· | ···················· |
| T | 1038 hCoV-19/England/QEUH-B6987B/2020\|EPI_ISL_661524\|20 | ···················· | ···················· | ····················· | ···················· | ···················· | ···················· |
| T | 1039 hCoV-19/England/QEUH-B6700C/2020\|EPI_ISL_661525\|20 | ···················· | ···················· | ····················· | ···················· | ···················· | ···················· |
| T | 1040 hCoV-19/England/QEUH-B692D7/2020\|EPI_ISL_661526\|20 | ···················· | ···················· | ····················· | ···················· | ···················· | ···················· |
| T | 1041 hCoV-19/England/QEUH-B6976F/2020\|EPI_ISL_661527\|20 | ···················· | ···················· | ····················· | ···················· | ···················· | ···················· |
| T | 1042 hCoV-19/England/QEUH-B67770/2020\|EPI_ISL_661528\|20 | ···················· | ···················· | ····················· | ···················· | ···T················ | ···················· |
| T | 1043 hCoV-19/England/QEUH-B68241/2020\|EPI_ISL_661529\|20 | ···················· | ···················· | ····················· | ···················· | ···················· | ···················· |
| T | 1044 hCoV-19/England/QEUH-B66586/2020\|EPI_ISL_661530\|20 | ···················· | ···················· | ····················· | ···················· | ···················· | ···················· |
| T | 1045 hCoV-19/England/QEUH-B683D5/2020\|EPI_ISL_661531\|20 | ···················· | ···················· | ····················· | ···················· | ···················· | ···················· |
| T | 1046 hCoV-19/England/QEUH-B69A84/2020\|EPI_ISL_661532\|20 | ···················· | ···················· | ····················· | ···················· | ···················· | ···················· |
| T | 1047 hCoV-19/England/QEUH-B6834E/2020\|EPI_ISL_661533\|20 | ···················· | ·A·················· | ····················· | ···················· | ···················· | ···················· |
| T | 1048 hCoV-19/England/QEUH-B67655/2020\|EPI_ISL_661534\|20 | ···················· | ···················· | ····················· | ···················· | ···················· | ···················· |
| T | 1049 hCoV-19/England/QEUH-B68C07/2020\|EPI_ISL_661535\|20 | ···················· | ···················· | ····················· | ···················· | ···················· | ···················· |
| T | 1050 hCoV-19/England/QEUH-B6C5AE/2020\|EPI_ISL_661537\|20 | ···················· | ···················· | ····················· | ···················· | ···················· | ···················· |
| T | 1051 hCoV-19/England/QEUH-B677DA/2020\|EPI_ISL_661539\|20 | ···················· | ···················· | ····················· | ···················· | ···················· | ···················· |
| T | 1052 hCoV-19/England/QEUH-B69AB1/2020\|EPI_ISL_661540\|20 | ···················· | ···················· | ····················· | ···················· | ···················· | ···················· |
| T | 1053 hCoV-19/England/QEUH-B6643E/2020\|EPI_ISL_661541\|20 | ···················· | ···················· | ····················· | ···················· | ···················· | ···················· |
| T | 1054 hCoV-19/England/QEUH-B66568/2020\|EPI_ISL_661542\|20 | ···················· | ···················· | ····················· | ···················· | ···················· | ···················· |
| T | 1055 hCoV-19/England/QEUH-B68469/2020\|EPI_ISL_661544\|20 | ···················· | ···················· | ····················· | ···················· | ···················· | ···················· |
| T | 1056 hCoV-19/England/QEUH-B6935C/2020\|EPI_ISL_661545\|20 | ···················· | ···················· | ····················· | ···················· | ···················· | ···················· |
| T | 1057 hCoV-19/England/QEUH-B69A66/2020\|EPI_ISL_661548\|20 | ···················· | ···················· | ····················· | ···················· | ···················· | ···················· |
| T | 1058 hCoV-19/England/QEUH-B68478/2020\|EPI_ISL_661549\|20 | ···················· | ···················· | ····················· | ···················· | ···················· | ···················· |
| T | 1059 hCoV-19/England/QEUH-B66FD3/2020\|EPI_ISL_661550\|20 | ···················· | ···················· | ····················· | ···················· | ···················· | ···················· |
| T | 1060 hCoV-19/England/QEUH-B679A7/2020\|EPI_ISL_661552\|20 | ···················· | ·A·················· | ····················· | ···················· | ···················· | ···················· |
| T | 1061 hCoV-19/England/QEUH-B6C67E/2020\|EPI_ISL_661554\|20 | ···················· | ···················· | ····················· | ···················· | ···················· | ···················· |
| T | 1062 hCoV-19/England/QEUH-B67BA1/2020\|EPI_ISL_661555\|20 | ···················· | ···················· | ····················· | ···················· | ···················· | ···················· |
| T | 1063 hCoV-19/England/QEUH-B67B74/2020\|EPI_ISL_661556\|20 | ···················· | ···················· | ····················· | ···················· | ···················· | ···················· |
| T | 1064 hCoV-19/England/QEUH-B66595/2020\|EPI_ISL_661557\|20 | ···················· | ···················· | ····················· | ···················· | ···················· | ···················· |
| T | 1065 hCoV-19/England/QEUH-B6C535/2020\|EPI_ISL_661558\|20 | ···················· | ···················· | ····················· | ···················· | ···················· | ···················· |
| T | 1066 hCoV-19/England/QEUH-B699D2/2020\|EPI_ISL_661559\|20 | ···················· | ···················· | ····················· | ···················· | ···················· | ···················· |
| T | 1067 hCoV-19/England/QEUH-B681F9/2020\|EPI_ISL_661560\|20 | ···················· | ···················· | ····················· | ···················· | ···················· | ···················· |
| T | 1068 hCoV-19/England/QEUH-B6979C/2020\|EPI_ISL_661561\|20 | ···················· | ···················· | ····················· | ···················· | ···················· | ···················· |
| T | 1069 hCoV-19/England/QEUH-B6637D/2020\|EPI_ISL_661562\|20 | ···················· | ···················· | ····················· | ···················· | ···················· | ···················· |
| T | 1070 hCoV-19/England/QEUH-B69301/2020\|EPI_ISL_661563\|20 | ···················· | ···················· | ····················· | ···················· | ···················· | ···················· |
| T | 1071 hCoV-19/England/QEUH-AEFDE8/2020\|EPI_ISL_662830\|20 | ···················· | ···················· | ····················· | ···················· | ···················· | ···················· |
| T | 1072 hCoV-19/England/QEUH-AF471F/2020\|EPI_ISL_662831\|20 | ···················· | ···················· | ····················· | ···················· | ···················· | ···················· |
| T | 1073 hCoV-19/England/QEUH-AF480D/2020\|EPI_ISL_662833\|20 | ···················· | ···················· | ····················· | ···················· | ···················· | ···················· |
| T | 1074 hCoV-19/England/QEUH-AEFBC0/2020\|EPI_ISL_662834\|20 | ···················· | ···················· | ····················· | ···················· | ···················· | ···················· |
| T | 1075 hCoV-19/England/QEUH-AF04EF/2020\|EPI_ISL_662835\|20 | ···················· | ···················· | ····················· | ···················· | ···················· | ···················· |
| T | 1076 hCoV-19/England/QEUH-AF28C3/2020\|EPI_ISL_662840\|20 | ···················· | ···················· | ····················· | ···················· | ···················· | ···················· |
| T | 1077 hCoV-19/England/QEUH-AF2896/2020\|EPI_ISL_662843\|20 | ···················· | ···················· | ····················· | ···················· | ···················· | ···················· |
| T | 1078 hCoV-19/England/QEUH-AEFA69/2020\|EPI_ISL_662844\|20 | ···················· | ···················· | ····················· | ···················· | ···················· | ···················· |
| T | 1079 hCoV-19/England/QEUH-AF00AF/2020\|EPI_ISL_662846\|20 | ···················· | ···················· | ····················· | ···················· | ···················· | ···················· |
| T | 1080 hCoV-19/England/QEUH-AF0740/2020\|EPI_ISL_662847\|20 | ···················· | ···················· | ····················· | ···················· | ···················· | ···················· |
| T | 1081 hCoV-19/England/QEUH-AF483A/2020\|EPI_ISL_662848\|20 | ···················· | ···················· | ····················· | ···················· | ···················· | ···················· |
| T | 1082 hCoV-19/England/QEUH-AF27A8/2020\|EPI_ISL_662850\|20 | ···················· | ···················· | ····················· | ···················· | ···················· | ···················· |
| T | 1083 hCoV-19/England/QEUH-AF4849/2020\|EPI_ISL_662851\|20 | ···················· | ···················· | ····················· | ···················· | ···················· | ···················· |
| T | 1084 hCoV-19/England/QEUH-AF0C8D/2020\|EPI_ISL_662853\|20 | ···················· | ···················· | ····················· | ···················· | ···················· | ···················· |
| T | 1085 hCoV-19/England/QEUH-AF0054/2020\|EPI_ISL_662856\|20 | ···················· | ···················· | ····················· | ···················· | ···················· | ···················· |
| T | 1086 hCoV-19/England/QEUH-AF0A0B/2020\|EPI_ISL_662857\|20 | ···················· | ···················· | ····················· | ···················· | ···················· | ···················· |
| T | 1087 hCoV-19/England/QEUH-AF0C50/2020\|EPI_ISL_662860\|20 | ···················· | ···················· | ····················· | ···················· | ···················· | ···················· |
| T | 1088 hCoV-19/England/QEUH-AF078C/2020\|EPI_ISL_662861\|20 | ···················· | ·A·················· | ····················· | ···················· | ···················· | ··A················· |
| T | 1089 hCoV-19/England/QEUH-AF4700/2020\|EPI_ISL_662862\|20 | ···················· | ···················· | ····················· | ···················· | ···················· | ···················· |
| T | 1090 hCoV-19/England/QEUH-AF0FA2/2020\|EPI_ISL_662863\|20 | ···················· | ···················· | ····················· | ···················· | ···················· | ···················· |
| T | 1091 hCoV-19/England/QEUH-AEFE03/2020\|EPI_ISL_662864\|20 | ···················· | ···················· | ····················· | ···················· | ···················· | ···················· |
| T | 1092 hCoV-19/England/QEUH-AEFFA6/2020\|EPI_ISL_662865\|20 | ···················· | ·A·················· | ····················· | ···················· | ···················· | ··A················· |
| T | 1093 hCoV-19/England/QEUH-AF46F4/2020\|EPI_ISL_662866\|20 | ···················· | ···················· | ····················· | ···················· | ···················· | ···················· |
| T | 1094 hCoV-19/England/QEUH-AF06DA/2020\|EPI_ISL_662867\|20 | ···················· | ···················· | ····················· | ···················· | ···················· | ···················· |
| T | 1095 hCoV-19/England/QEUH-AEFFB5/2020\|EPI_ISL_662868\|20 | ···················· | ···················· | ····················· | ···················· | ···················· | ···················· |
| T | 1096 hCoV-19/England/QEUH-AF2878/2020\|EPI_ISL_662870\|20 | ···················· | ···················· | ····················· | ···················· | ···················· | ···················· |
| T | 1097 hCoV-19/England/QEUH-AF0C6F/2020\|EPI_ISL_662871\|20 | ···················· | ·A·················· | ····················· | ···················· | ···················· | ···················· |
| T | 1098 hCoV-19/England/QEUH-AF0CAB/2020\|EPI_ISL_662872\|20 | ···················· | ···················· | ····················· | ···················· | ···················· | ···················· |
| T | 1099 hCoV-19/England/QEUH-AF0661/2020\|EPI_ISL_662873\|20 | ···················· | ···················· | ····················· | ···················· | ···················· | ···················· |
| T | 1100 hCoV-19/England/QEUH-AEFBDF/2020\|EPI_ISL_662875\|20 | ···················· | ···················· | ····················· | ···················· | ···················· | ···················· |

| | | | | | | | |
|---|---|---|---|---|---|---|---|
| T | 1101 hCoV-19/England/QEUH-AF0133/2020\|EPI_ISL_662876\|20 | ···················· | ···················· | ····················· | ···················· | ···················· | ···················· |
| T | 1102 hCoV-19/England/QEUH-AF0519/2020\|EPI_ISL_662878\|20 | ···················· | ···················· | ····················· | ···················· | ···················· | ···················· |
| T | 1103 hCoV-19/England/QEUH-AF763D/2020\|EPI_ISL_662879\|20 | ···················· | ···················· | ····················· | ···················· | ···················· | ···················· |
| T | 1104 hCoV-19/England/QEUH-AEF9E4/2020\|EPI_ISL_662880\|20 | ···················· | ···················· | ····················· | ···················· | ···················· | ···················· |
| T | 1105 hCoV-19/England/QEUH-AF47E2/2020\|EPI_ISL_662881\|20 | ···················· | ·A·················· | ····················· | ···················· | ···················· | ···················· |
| T | 1106 hCoV-19/England/QEUH-AEFD60/2020\|EPI_ISL_662882\|20 | ···················· | ···················· | ····················· | ···················· | ···················· | ···················· |
| T | 1107 hCoV-19/England/QEUH-AF2553/2020\|EPI_ISL_662883\|20 | ···················· | ·A·················· | ····················· | ···················· | ···················· | ··A················· |
| T | 1108 hCoV-19/England/QEUH-AF1026/2020\|EPI_ISL_662884\|20 | ···················· | ···················· | ····················· | ···················· | ···················· | ···················· |
| T | 1109 hCoV-19/England/QEUH-AEFFC4/2020\|EPI_ISL_662886\|20 | ···················· | ···················· | ····················· | ···················· | ···················· | ···················· |
| T | 1110 hCoV-19/England/QEUH-AEFE21/2020\|EPI_ISL_662887\|20 | ···················· | ···················· | ····················· | ···················· | ···················· | ···················· |
| T | 1111 hCoV-19/England/QEUH-AF1062/2020\|EPI_ISL_662888\|20 | ···················· | ···················· | ····················· | ···················· | ···················· | ···················· |
| T | 1112 hCoV-19/England/QEUH-AF0F93/2020\|EPI_ISL_662889\|20 | ···················· | ···················· | ····················· | ···················· | ···················· | ···················· |
| T | 1113 hCoV-19/England/QEUH-AF48D0/2020\|EPI_ISL_662890\|20 | ···················· | ···················· | ····················· | ···················· | ···················· | ···················· |
| T | 1114 hCoV-19/England/QEUH-AF4A25/2020\|EPI_ISL_662891\|20 | ···················· | ···················· | ····················· | ···················· | ···················· | ···················· |
| T | 1115 hCoV-19/England/QEUH-AF4D68/2020\|EPI_ISL_662892\|20 | ···················· | ···················· | ····················· | ···················· | ···················· | ···················· |
| T | 1116 hCoV-19/England/QEUH-AF4D0E/2020\|EPI_ISL_662893\|20 | ···················· | ···················· | ····················· | ···················· | ···················· | ···················· |
| T | 1118 hCoV-19/England/QEUH-AF559C/2020\|EPI_ISL_662900\|20 | ···················· | ···················· | ····················· | ···················· | ···················· | ···················· |
| T | 1119 hCoV-19/England/QEUH-AF3DC3/2020\|EPI_ISL_662901\|20 | ···················· | ···················· | ····················· | ···················· | ···················· | ···················· |
| T | 1120 hCoV-19/England/QEUH-AF4DFF/2020\|EPI_ISL_662902\|20 | ···················· | ···················· | ····················· | ···················· | ···················· | ···················· |
| T | 1121 hCoV-19/England/QEUH-AF68ED/2020\|EPI_ISL_662903\|20 | ···················· | ···················· | ····················· | ···················· | ···················· | ···················· |
| T | 1122 hCoV-19/England/QEUH-AF565D/2020\|EPI_ISL_662904\|20 | ···················· | ···················· | ····················· | ···················· | ···················· | ···················· |
| T | 1123 hCoV-19/England/QEUH-AF58A2/2020\|EPI_ISL_662905\|20 | ···················· | ···················· | ····················· | ···················· | ···················· | ···················· |
| T | 1124 hCoV-19/England/QEUH-AF4C4D/2020\|EPI_ISL_662906\|20 | ···················· | ···················· | ····················· | ···················· | ···················· | ···················· |
| T | 1125 hCoV-19/England/QEUH-AF4B7D/2020\|EPI_ISL_662907\|20 | ···················· | ···················· | ····················· | ···················· | ···················· | ···················· |
| T | 1126 hCoV-19/England/QEUH-AF57F0/2020\|EPI_ISL_662908\|20 | ···················· | ···················· | ····················· | ···················· | ···················· | ···················· |
| T | 1127 hCoV-19/England/QEUH-AF3E1B/2020\|EPI_ISL_662909\|20 | ···················· | ·A·················· | ····················· | ···················· | ···················· | ···················· |
| T | 1128 hCoV-19/England/QEUH-AF5417/2020\|EPI_ISL_662910\|20 | ···················· | ···················· | ····················· | ···················· | ···················· | ···················· |
| T | 1129 hCoV-19/England/QEUH-AF58FD/2020\|EPI_ISL_662911\|20 | ···················· | ···················· | ····················· | ···················· | ···················· | ···················· |
| T | 1130 hCoV-19/England/QEUH-AF6926/2020\|EPI_ISL_662912\|20 | ···················· | ·A·················· | ····················· | ···················· | ···················· | ···················· |
| T | 1131 hCoV-19/England/QEUH-AF4DE0/2020\|EPI_ISL_662913\|20 | ···················· | ···················· | ····················· | ···················· | ···················· | ···················· |
| T | 1132 hCoV-19/England/QEUH-AF6698/2020\|EPI_ISL_662914\|20 | ···················· | ···················· | ····················· | ···················· | ···················· | ···················· |
| T | 1133 hCoV-19/England/QEUH-AF56F3/2020\|EPI_ISL_662915\|20 | ···················· | ···················· | ····················· | ···················· | ···················· | ···················· |
| T | 1134 hCoV-19/England/QEUH-AF5444/2020\|EPI_ISL_662916\|20 | ···················· | ···················· | ····················· | ···················· | ···················· | ···················· |
| T | 1135 hCoV-19/England/QEUH-AF58EE/2020\|EPI_ISL_662917\|20 | ···················· | ·A·················· | ····················· | ···················· | ···················· | ···················· |
| T | 1136 hCoV-19/England/QEUH-AF49FB/2020\|EPI_ISL_662918\|20 | ···················· | ···················· | ····················· | ···················· | ···················· | ···················· |
| T | 1137 hCoV-19/England/QEUH-AF61C4/2020\|EPI_ISL_662919\|20 | ···················· | ···················· | ····················· | ···················· | ···················· | ···················· |
| T | 1138 hCoV-19/England/QEUH-AF6953/2020\|EPI_ISL_662921\|20 | ···················· | ···················· | ····················· | ···················· | ···················· | ···················· |
| T | 1139 hCoV-19/England/QEUH-AF5787/2020\|EPI_ISL_662922\|20 | ···················· | ···················· | ····················· | ···················· | ···················· | ···················· |
| T | 1140 hCoV-19/England/QEUH-AF3D3C/2020\|EPI_ISL_662923\|20 | ···················· | ···················· | ····················· | ···················· | ···················· | ···················· |
| T | 1141 hCoV-19/England/QEUH-AF673B/2020\|EPI_ISL_662924\|20 | ···················· | ···················· | ····················· | ···················· | ···················· | ···················· |
| T | 1142 hCoV-19/England/QEUH-AF5189/2020\|EPI_ISL_662925\|20 | ···················· | ···················· | ····················· | ···················· | ···················· | ···················· |
| T | 1143 hCoV-19/England/QEUH-AF5611/2020\|EPI_ISL_662926\|20 | ···················· | ···················· | ····················· | ···················· | ···················· | ···················· |
| T | 1144 hCoV-19/England/QEUH-AF4A43/2020\|EPI_ISL_662927\|20 | ···················· | ···················· | ····················· | ···················· | ···················· | ···················· |
| T | 1145 hCoV-19/England/QEUH-AF5699/2020\|EPI_ISL_662928\|20 | ···················· | ·A·················· | ····················· | ···················· | ···················· | ··A················· |
| T | 1146 hCoV-19/England/QEUH-AF5101/2020\|EPI_ISL_662929\|20 | ···················· | ···················· | ····················· | ···················· | ···················· | ···················· |
| T | 1147 hCoV-19/England/LIVE-DBD15D/2020\|EPI_ISL_665128\|20 | ···················· | ···················· | ····················· | ···················· | ···················· | ···················· |
| T | 1148 hCoV-19/England/LIVE-DBD1F3/2020\|EPI_ISL_665129\|20 | ···················· | ···················· | ····················· | ···················· | ···················· | ···················· |
| T | 1149 hCoV-19/England/LIVE-DBD22D/2020\|EPI_ISL_665130\|20 | ···················· | ···················· | ····················· | ···················· | ···················· | ···················· |
| T | 1150 hCoV-19/England/LIVE-DBD30C/2020\|EPI_ISL_665131\|20 | ···················· | ···················· | ····················· | ···················· | ···················· | ···················· |
| T | 1151 hCoV-19/England/LIVE-DBD384/2020\|EPI_ISL_665132\|20 | ···················· | ···················· | ····················· | ···················· | ···················· | ···················· |
| T | 1152 hCoV-19/England/LIVE-DBD3A2/2020\|EPI_ISL_665133\|20 | ···················· | ···················· | ····················· | ···················· | ···················· | ···················· |
| T | 1154 hCoV-19/England/LIVE-DBD603/2020\|EPI_ISL_665135\|20 | ···················· | ···················· | ····················· | ···················· | ···················· | ···················· |
| T | 1159 hCoV-19/England/LIVE-DC1208/2020\|EPI_ISL_665140\|20 | ···················· | ···················· | ····················· | ···················· | ···················· | ···················· |
| T | 1162 hCoV-19/England/LOND-124F158/2020\|EPI_ISL_665143\|2 | ···················· | ···················· | ····················· | ···················· | ···················· | ···················· |
| T | 1163 hCoV-19/England/LOND-124F167/2020\|EPI_ISL_665144\|2 | ···················· | ···················· | ····················· | ···················· | ···················· | ···················· |
| T | 1164 hCoV-19/England/LOND-124F1B2/2020\|EPI_ISL_665145\|2 | ···················· | ···················· | ····················· | ···················· | ···················· | ···················· |
| T | 1167 hCoV-19/England/LOND-124F282/2020\|EPI_ISL_665148\|2 | ···················· | ···················· | ····················· | ···················· | ···················· | ···················· |
| T | 1168 hCoV-19/England/LOND-124F2DD/2020\|EPI_ISL_665149\|2 | ···················· | ···················· | ····················· | ···················· | ···NN··············· | ···················· |
| T | 1170 hCoV-19/England/LOND-124F3AD/2020\|EPI_ISL_665151\|2 | ···················· | ···················· | ····················· | ···················· | ···················· | ···················· |
| T | 1171 hCoV-19/England/LOND-124F413/2020\|EPI_ISL_665152\|2 | ···················· | ·A·················· | ····················· | ···················· | ···················· | ···················· |
| T | 1172 hCoV-19/England/LOND-124F46E/2020\|EPI_ISL_665153\|2 | ···················· | ···················· | ····················· | ···················· | ···················· | ···················· |
| T | 1173 hCoV-19/England/LOND-124F47D/2020\|EPI_ISL_665154\|2 | ···················· | ···················· | ····················· | ···················· | ···················· | ···················· |
| T | 1174 hCoV-19/England/LOND-124F48C/2020\|EPI_ISL_665155\|2 | ···················· | ···················· | ····················· | ···················· | ···················· | ···················· |
| T | 1175 hCoV-19/England/LOND-124F49B/2020\|EPI_ISL_665156\|2 | ···················· | ···················· | ····················· | ···················· | ···················· | ···················· |
| T | 1176 hCoV-19/England/LOND-124F4AA/2020\|EPI_ISL_665157\|2 | ···················· | ·A·················· | ····················· | ···················· | ···················· | ··A················· |
| T | 1177 hCoV-19/England/LOND-124F4D7/2020\|EPI_ISL_665158\|2 | ···················· | ···················· | ····················· | ···················· | ···················· | ···················· |
| T | 1178 hCoV-19/England/LOND-124F4E6/2020\|EPI_ISL_665159\|2 | ···················· | ···················· | ····················· | ···················· | ···················· | ···················· |
| T | 1179 hCoV-19/England/LOND-124F510/2020\|EPI_ISL_665160\|2 | ···················· | ···················· | ····················· | ···················· | ···················· | ···················· |
| T | 1181 hCoV-19/England/LOND-124F659/2020\|EPI_ISL_665162\|2 | ···················· | ···················· | ····················· | ···················· | ···················· | ···················· |
| T | 1182 hCoV-19/England/LOND-124F668/2020\|EPI_ISL_665163\|2 | ···················· | ···················· | ····················· | ···················· | ···················· | ···················· |
| T | 1183 hCoV-19/England/LOND-124F6C2/2020\|EPI_ISL_665164\|2 | ···················· | ···················· | ····················· | ···················· | ···················· | ···················· |
| T | 1184 hCoV-19/England/LOND-124F6D1/2020\|EPI_ISL_665165\|2 | ···················· | ···················· | ····················· | ···················· | ···················· | ···················· |
| T | 1185 hCoV-19/England/LOND-124F6FF/2020\|EPI_ISL_665166\|2 | ···················· | ···················· | ····················· | ···················· | ···················· | ···················· |
| T | 1186 hCoV-19/England/LOND-124F71A/2020\|EPI_ISL_665167\|2 | ···················· | ···················· | ····················· | ···················· | ···················· | ···················· |
| T | 1187 hCoV-19/England/LOND-124F738/2020\|EPI_ISL_665168\|2 | ···················· | ···················· | ····················· | ···················· | ···················· | ···················· |
| T | 1188 hCoV-19/England/LOND-124F756/2020\|EPI_ISL_665169\|2 | ···················· | ···················· | ····················· | ···················· | ···················· | ···················· |
| T | 1189 hCoV-19/England/LOND-124F765/2020\|EPI_ISL_665170\|2 | ···················· | ···················· | ····················· | ···················· | ···················· | ···················· |
| T | 1190 hCoV-19/England/LOND-124F792/2020\|EPI_ISL_665171\|2 | ···················· | ···················· | ····················· | ···················· | ···················· | ···················· |
| T | 1192 hCoV-19/England/LOND-124F7B0/2020\|EPI_ISL_665173\|2 | ···················· | ···················· | ····················· | ···················· | ···················· | ···················· |
| T | 1195 hCoV-19/England/LOND-124F880/2020\|EPI_ISL_665176\|2 | ···················· | ···················· | ····················· | ···················· | ···················· | ···················· |

T 1197 hCoV-19/England/LOND-124F923/2020|EPI_ISL_665178|2 ···················· ···················· ····················· ···················· ···················· ····················
T 1199 hCoV-19/England/LOND-124F98D/2020|EPI_ISL_665180|2 ···················· ···················· ····················· ···················· ···················· ····················
T 1200 hCoV-19/England/LOND-124F9AB/2020|EPI_ISL_665181|2 ···················· ···················· ····················· ···················· ···················· ····················
T 1201 hCoV-19/England/LOND-124FB0F/2020|EPI_ISL_665182|2 ···················· ···················· ····················· ···················· ···················· ····················
T 1202 hCoV-19/England/LOND-124FB1E/2020|EPI_ISL_665183|2 ···················· ···················· ····················· ···················· ···················· ····················
T 1203 hCoV-19/England/LOND-124FB2D/2020|EPI_ISL_665184|2 ···················· ···················· ····················· ···················· ···················· ····················
T 1204 hCoV-19/England/LOND-124FB3C/2020|EPI_ISL_665185|2 ···················· ···················· ····················· ···················· ···················· ····················
T 1205 hCoV-19/England/LOND-124FB5A/2020|EPI_ISL_665186|2 ···················· ···················· ····················· ···················· ···················· ····················
T 1206 hCoV-19/England/LOND-124FB78/2020|EPI_ISL_665187|2 ···················· ···················· ····················· ···················· ···················· ····················
T 1207 hCoV-19/England/LOND-124FBA5/2020|EPI_ISL_665188|2 ···················· ···················· ····················· ···················· ···················· ····················
T 1208 hCoV-19/England/LOND-124FBC3/2020|EPI_ISL_665189|2 ···················· ···················· ····················· ···················· ···················· ····················
T 1209 hCoV-19/England/LOND-124FE60/2020|EPI_ISL_665190|2 ···················· ···················· ····················· ···················· ···················· ····················
T 1210 hCoV-19/England/LOND-124FFF4/2020|EPI_ISL_665191|2 ···················· ···················· ····················· ···················· ···················· ····················
T 1211 hCoV-19/England/LOND-1250039/2020|EPI_ISL_665192|2 ···················· ···················· ····················· ···················· ···················· ····················
T 1212 hCoV-19/England/LOND-1250093/2020|EPI_ISL_665193|2 ···················· ···················· ····················· ···················· ···················· ····················
T 1213 hCoV-19/England/LOND-12500B1/2020|EPI_ISL_665194|2 ···················· ···················· ····················· ···················· ···················· ····················
T 1214 hCoV-19/England/LOND-12500FD/2020|EPI_ISL_665195|2 ···················· ···················· ····················· ···················· ···················· ····················
T 1215 hCoV-19/England/LOND-1250118/2020|EPI_ISL_665196|2 ···················· ···················· ····················· ···················· ···················· ····················
T 1216 hCoV-19/England/LOND-1250154/2020|EPI_ISL_665197|2 ···················· ···················· ····················· ···················· ···················· ····················
T 1217 hCoV-19/England/LOND-1250163/2020|EPI_ISL_665198|2 ···················· ···················· ····················· ···················· ···················· ····················
T 1218 hCoV-19/England/LOND-1250190/2020|EPI_ISL_665199|2 ···················· ···················· ····················· ···················· ···················· ····················
T 1220 hCoV-19/Wales/PHWC-48C703/2020|EPI_ISL_666460|2020 ···················· ···················· ····················· ···················· ···················· ····················
T 1221 hCoV-19/Wales/PHWC-48C721/2020|EPI_ISL_666461|2020 ···················· ···················· ····················· ···················· ···················· ····················
T 1222 hCoV-19/Wales/PHWC-48C730/2020|EPI_ISL_666462|2020 ···················· ···················· ····················· ···················· ···················· ····················
T 1223 hCoV-19/Wales/PHWC-48C74F/2020|EPI_ISL_666463|2020 ···················· ···················· ····················· ···················· ···················· ····················
T 1224 hCoV-19/Wales/PHWC-48C75E/2020|EPI_ISL_666464|2020 ···················· ···················· ····················· ···················· ···················· ····················
T 1225 hCoV-19/Wales/PHWC-48C78B/2020|EPI_ISL_666465|2020 ···················· ···················· ····················· ···················· ···················· ····················
T 1226 hCoV-19/Wales/PHWC-48C7A9/2020|EPI_ISL_666466|2020 ···················· ···················· ····················· ···················· ···················· ····················
T 1227 hCoV-19/Wales/PHWC-48C7B8/2020|EPI_ISL_666467|2020 ···················· ···················· ····················· ···················· ···················· ····················
T 1228 hCoV-19/Wales/PHWC-48C7C7/2020|EPI_ISL_666468|2020 ···················· ···················· ····················· ···················· NN·················· ····················
T 1229 hCoV-19/Wales/PHWC-48C7D6/2020|EPI_ISL_666469|2020 ···················· ···················· ····················· ···················· ···················· ····················
T 1230 hCoV-19/Wales/PHWC-48C7E5/2020|EPI_ISL_666470|2020 ···················· ···················· ····················· ···················· ···················· ····················
T 1231 hCoV-19/Wales/PHWC-48C7F4/2020|EPI_ISL_666471|2020 ···················· ···················· ····················· ···················· ···················· ····················
T 1232 hCoV-19/Wales/PHWC-48C800/2020|EPI_ISL_666472|2020 ···················· ···················· ····················· ···················· ···················· ····················
T 1233 hCoV-19/Wales/PHWC-48C81F/2020|EPI_ISL_666473|2020 ···················· ···················· ····················· ···················· ···················· ····················
T 1234 hCoV-19/Wales/PHWC-48C82E/2020|EPI_ISL_666474|2020 ···················· ···················· ····················· ···················· ···················· ····················
T 1235 hCoV-19/Wales/PHWC-48C83D/2020|EPI_ISL_666475|2020 ···················· ···················· ····················· ···················· ···················· ····················
T 1236 hCoV-19/Wales/PHWC-48C84C/2020|EPI_ISL_666476|2020 ···················· ·R·················· ····················· ···················· ···················· ····················
T 1237 hCoV-19/Wales/PHWC-48C86A/2020|EPI_ISL_666477|2020 ···················· ···················· ····················· ···················· ···················· ····················
T 1238 hCoV-19/Wales/PHWC-48C879/2020|EPI_ISL_666478|2020 ···················· ···················· ····················· ···················· ···················· ····················
T 1240 hCoV-19/Wales/PHWC-48C8B5/2020|EPI_ISL_666480|2020 ···················· ···················· ····················· ···················· ···················· ····················
T 1241 hCoV-19/Wales/PHWC-48C8C4/2020|EPI_ISL_666481|2020 ···················· ···················· ····················· ···················· ···················· ····················
T 1242 hCoV-19/Wales/PHWC-48C8D3/2020|EPI_ISL_666482|2020 ···················· ···················· ····················· ···················· ···················· ····················
T 1243 hCoV-19/Wales/PHWC-48C8E2/2020|EPI_ISL_666483|2020 ···················· ···················· ····················· ···················· ···················· ····················
T 1244 hCoV-19/Wales/PHWC-48C90D/2020|EPI_ISL_666484|2020 ···················· ···················· ····················· ···················· ···················· ····················
T 1245 hCoV-19/Wales/PHWC-48C91C/2020|EPI_ISL_666485|2020 ···················· ···················· ····················· ···················· ···················· ····················
T 1246 hCoV-19/Wales/PHWC-48C92B/2020|EPI_ISL_666486|2020 ···················· ···················· ····················· ···················· ···················· ····················
T 1247 hCoV-19/Wales/PHWC-48C949/2020|EPI_ISL_666487|2020 ···················· ···················· ····················· ···················· ·NN················· ····················
T 1248 hCoV-19/Wales/PHWC-48C976/2020|EPI_ISL_666488|2020 ···················· ···················· ····················· ···················· ···················· ····················
T 1249 hCoV-19/Wales/PHWC-48C985/2020|EPI_ISL_666489|2020 ···················· ···················· ····················· ···················· ···················· ····················
T 1250 hCoV-19/Wales/PHWC-48C9B2/2020|EPI_ISL_666490|2020 ···················· ···················· ····················· ···················· ···················· ····················
T 1251 hCoV-19/Wales/PHWC-48C9C1/2020|EPI_ISL_666491|2020 ···················· ···················· ····················· ···················· NNN················· ···················· 1
T 1252 hCoV-19/Wales/PHWC-48C9EF/2020|EPI_ISL_666492|2020 ···················· ···················· ····················· ···················· ···················· ····················
T 1253 hCoV-19/Wales/PHWC-48C9FE/2020|EPI_ISL_666493|2020 ···················· ···················· ····················· ···················· ···················· ····················
T 1254 hCoV-19/Wales/PHWC-48CA0A/2020|EPI_ISL_666494|2020 ···················· ···················· ····················· ···················· ···················· ····················
T 1256 hCoV-19/Wales/PHWC-48CA28/2020|EPI_ISL_666496|2020 ···················· ···················· ····················· ···················· ···················· ····················
T 1257 hCoV-19/Wales/PHWC-48CA37/2020|EPI_ISL_666497|2020 ···················· ···················· ····················· ···················· ···················· ····················
T 1258 hCoV-19/Wales/PHWC-48CA55/2020|EPI_ISL_666498|2020 ···················· ···················· ····················· ···················· ···················· ····················
T 1259 hCoV-19/Wales/PHWC-48CA64/2020|EPI_ISL_666499|2020 ···················· ···················· ····················· ···················· ···················· ····················
T 1260 hCoV-19/Australia/NSW1189/2020|EPI_ISL_667790|2020 ···················· ···················· ····················· ···················· ···················· ····················
T 1261 hCoV-19/Australia/NSW1190/2020|EPI_ISL_667791|2020 ···················· ···················· ····················· ···················· ···················· ····················
T 1262 hCoV-19/Australia/NSW1193/2020|EPI_ISL_667793|2020 ···················· ···················· ····················· ···················· ···················· ····················
T 1263 hCoV-19/Australia/NSW1194/2020|EPI_ISL_667794|2020 ···················· ·A·················· ····················· ···················· ·········T·········· ···A················
T 1264 hCoV-19/Australia/NSW1195/2020|EPI_ISL_667795|2020 ···················· ···················· ····················· ···················· ···················· ····················
T 1265 hCoV-19/Australia/NSW1196/2020|EPI_ISL_667796|2020 ···················· ·A·················· ····················· ···················· ···················· ···A················
T 1266 hCoV-19/Australia/NSW1197/2020|EPI_ISL_667797|2020 ···················· ···················· ····················· ···················· ···················· ····················
T 1267 hCoV-19/Australia/NSW1198/2020|EPI_ISL_667798|2020 ···················· ···················· ····················· ···················· ···················· ····················
T 1268 hCoV-19/Australia/NSW1199/2020|EPI_ISL_667799|2020 ···················· ···················· ····················· ···················· ···················· ····················
T 1269 hCoV-19/USA/CA-CZB-10048/2020|EPI_ISL_672200|2020- ···················· ···················· ····················· ···················· ···················· ····················
T 1270 hCoV-19/USA/CA-CZB-10064/2020|EPI_ISL_672201|2020- ···················· ···················· ····················· ···················· ···················· ····················
T 1271 hCoV-19/USA/CA-CZB-10065/2020|EPI_ISL_672202|2020- ···················· ···················· ····················· ···················· ···················· ····················
T 1272 hCoV-19/USA/CA-CZB-10067/2020|EPI_ISL_672203|2020- ···················· ···················· ····················· ···················· ···················· ····················
T 1273 hCoV-19/USA/CA-CZB-10070/2020|EPI_ISL_672204|2020- ···················· ···················· ····················· ···················· ···················· ····················
T 1274 hCoV-19/USA/CA-CZB-10073/2020|EPI_ISL_672205|2020- ···················· ···················· ····················· ···················· ···················· ····················
T 1275 hCoV-19/USA/CA-CZB-10076/2020|EPI_ISL_672206|2020- ········Y··········· ·A·················· ····················· ···················· ···················· ····················
T 1276 hCoV-19/USA/CA-CZB-10077/2020|EPI_ISL_672207|2020- ···················· ·A·················· ····················· ···················· ···················· ····················
T 1277 hCoV-19/USA/CA-CZB-10082/2020|EPI_ISL_672208|2020- ···················· ···················· ····················· ···················· ···················· ····················
T 1278 hCoV-19/USA/CA-CZB-10085/2020|EPI_ISL_672209|2020- ···················· ···················· ····················· ···················· ···················· ····················
T 1279 hCoV-19/USA/CA-CZB-10091/2020|EPI_ISL_672210|2020- ···················· ···················· ····················· ···················· ···················· ····················
T 1280 hCoV-19/USA/CA-CZB-10096/2020|EPI_ISL_672211|2020- ···················· ···················· ····················· ···················· ···················· ····················

| | | | | | | | |
|---|---|---|---|---|---|---|---|
| T | 1281 hCoV-19/England/QEUH-B6840F/2020\|EPI_ISL_661505\|20 | ···················· | ···················· | ····················· | ···················· | ···················· | ···················· |
| T | 1282 hCoV-19/England/QEUH-B67B38/2020\|EPI_ISL_661506\|20 | ···················· | ···················· | ····················· | ···················· | ···················· | ···················· |
| T | 1283 hCoV-19/England/QEUH-B69B27/2020\|EPI_ISL_661507\|20 | ···················· | ···················· | ····················· | ···················· | ···················· | ···················· |
| T | 1284 hCoV-19/England/QEUH-B682F6/2020\|EPI_ISL_661508\|20 | ···················· | ···················· | ····················· | ···················· | ···················· | ···················· |
| T | 1285 hCoV-19/England/QEUH-B67707/2020\|EPI_ISL_661509\|20 | ···················· | ···················· | ····················· | ···················· | ···················· | ···················· |
| T | 1286 hCoV-19/England/QEUH-B67E3F/2020\|EPI_ISL_661564\|20 | ···················· | ·A·················· | ····················· | ···················· | ···················· | ···················· |
| T | 1287 hCoV-19/England/QEUH-B67A2C/2020\|EPI_ISL_661565\|20 | ···················· | ···················· | ····················· | ···················· | ···················· | ···················· |
| T | 1288 hCoV-19/England/QEUH-B662DA/2020\|EPI_ISL_661566\|20 | ···················· | ···················· | ····················· | ···················· | ···················· | ···················· |
| T | 1289 hCoV-19/England/QEUH-B68223/2020\|EPI_ISL_661567\|20 | ···················· | ···················· | ····················· | ···················· | ···················· | ···················· |
| T | 1290 hCoV-19/England/QEUH-B6936B/2020\|EPI_ISL_661568\|20 | ···················· | ···················· | ····················· | ···················· | ···················· | ···················· |
| T | 1291 hCoV-19/England/QEUH-B68487/2020\|EPI_ISL_661569\|20 | ···················· | ···················· | ····················· | ···················· | ···················· | ···················· |
| T | 1292 hCoV-19/England/QEUH-B69AC0/2020\|EPI_ISL_661571\|20 | ···················· | ···················· | ····················· | ···················· | ···················· | ···················· |
| T | 1293 hCoV-19/England/QEUH-B666A1/2020\|EPI_ISL_661572\|20 | ···················· | ···················· | ····················· | ···················· | ···················· | ···················· |
| T | 1294 hCoV-19/England/QEUH-B67C17/2020\|EPI_ISL_661573\|20 | ···················· | ···················· | ····················· | ···················· | ···················· | ···················· |
| T | 1295 hCoV-19/England/QEUH-B67B29/2020\|EPI_ISL_661574\|20 | ···················· | ·A·················· | ····················· | ···················· | ···················· | ···················· |
| T | 1296 hCoV-19/England/QEUH-B663B9/2020\|EPI_ISL_661575\|20 | ···················· | ···················· | ····················· | ···················· | ···················· | ···················· |
| T | 1297 hCoV-19/England/QEUH-B67673/2020\|EPI_ISL_661576\|20 | ···················· | ···················· | ····················· | ···················· | ···················· | ···················· |
| T | 1298 hCoV-19/England/QEUH-B676FB/2020\|EPI_ISL_661577\|20 | ···················· | ···················· | ····················· | ···················· | ····T··············· | ···················· |
| T | 1299 hCoV-19/England/QEUH-B689E2/2020\|EPI_ISL_661578\|20 | ···················· | ···················· | ····················· | ···················· | ···················· | ···················· |
| T | 1300 hCoV-19/England/QEUH-B68E4D/2020\|EPI_ISL_661579\|20 | ···················· | ···················· | ····················· | ···················· | ···················· | ···················· |
| T | 1301 hCoV-19/England/QEUH-B683E4/2020\|EPI_ISL_661580\|20 | ···················· | ···················· | ····················· | ···················· | ···················· | ···················· |
| T | 1302 hCoV-19/England/QEUH-B66225/2020\|EPI_ISL_661584\|20 | ···················· | ···················· | ····················· | ···················· | ···················· | ···················· |
| T | 1303 hCoV-19/England/QEUH-B6702A/2020\|EPI_ISL_661585\|20 | ···················· | ···················· | ····················· | ···················· | ···················· | ···················· |
| T | 1304 hCoV-19/England/QEUH-B68F1D/2020\|EPI_ISL_661586\|20 | ···················· | ···················· | ····················· | ···················· | ···················· | ···················· |
| T | 1305 hCoV-19/England/QEUH-B679B6/2020\|EPI_ISL_661587\|20 | ···················· | ·A·················· | ····················· | ···················· | ···················· | ···················· |
| T | 1306 hCoV-19/England/QEUH-B689C4/2020\|EPI_ISL_661591\|20 | ···················· | ···················· | ····················· | ···················· | ···················· | ···················· |
| T | 1307 hCoV-19/England/QEUH-B665A4/2020\|EPI_ISL_661592\|20 | ···················· | ···················· | ····················· | ···················· | ···················· | ···················· |
| T | 1308 hCoV-19/England/QEUH-B67C44/2020\|EPI_ISL_661593\|20 | ···················· | ···················· | ····················· | ···················· | ···················· | ···················· |
| T | 1309 hCoV-19/England/QEUH-B67C71/2020\|EPI_ISL_661595\|20 | ···················· | ···················· | ····················· | ···················· | ···················· | ···················· |
| T | 1310 hCoV-19/England/QEUH-B67E5D/2020\|EPI_ISL_661596\|20 | ···················· | ···················· | ····················· | ···················· | ···················· | ···················· |
| T | 1311 hCoV-19/England/QEUH-B6636E/2020\|EPI_ISL_661597\|20 | ···················· | ···················· | ····················· | ···················· | ···················· | ···················· |
| T | 1312 hCoV-19/England/QEUH-B69B63/2020\|EPI_ISL_661598\|20 | ···················· | ···················· | ····················· | ···················· | ···················· | ···················· |
| T | 1313 hCoV-19/England/QEUH-B699B4/2020\|EPI_ISL_661599\|20 | ···················· | ···················· | ····················· | ···················· | ···················· | ···················· |
| T | 1314 hCoV-19/England/QEUH-AF05BF/2020\|EPI_ISL_662811\|20 | ···················· | ···················· | ····················· | ···················· | ···················· | ···················· |
| T | 1315 hCoV-19/England/QEUH-AF09C2/2020\|EPI_ISL_662813\|20 | ···················· | ···················· | ····················· | ···················· | ···················· | ···················· |
| T | 1316 hCoV-19/England/QEUH-AF27E4/2020\|EPI_ISL_662814\|20 | ···················· | ·A·················· | ····················· | ···················· | ···················· | ···················· |
| T | 1317 hCoV-19/England/QEUH-AF01E8/2020\|EPI_ISL_662815\|20 | ···················· | ···················· | ····················· | ···················· | ···················· | ···················· |
| T | 1318 hCoV-19/England/QEUH-AF0C7E/2020\|EPI_ISL_662818\|20 | ···················· | ···················· | ····················· | ···················· | ···················· | ···················· |
| T | 1319 hCoV-19/England/QEUH-AF25CC/2020\|EPI_ISL_662819\|20 | ···················· | ·A·················· | ····················· | ···················· | ···················· | ···················· |
| T | 1320 hCoV-19/England/QEUH-AF472E/2020\|EPI_ISL_662823\|20 | ···················· | ···················· | ····················· | ···················· | ···················· | ···················· |
| T | 1321 hCoV-19/England/QEUH-AF050A/2020\|EPI_ISL_662824\|20 | ···················· | ···················· | ····················· | ···················· | ···················· | ···················· |
| T | 1322 hCoV-19/England/QEUH-AF27C6/2020\|EPI_ISL_662825\|20 | ···················· | ···················· | ····················· | ···················· | ···················· | ···················· |
| T | 1323 hCoV-19/England/QEUH-AF47F1/2020\|EPI_ISL_662826\|20 | ···················· | ···················· | ····················· | ···················· | ···················· | ···················· |
| T | 1324 hCoV-19/England/QEUH-AF0670/2020\|EPI_ISL_662829\|20 | ···················· | ···················· | ····················· | ···················· | ···················· | ···················· |
| T | 1325 hCoV-19/England/QEUH-AF46E5/2020\|EPI_ISL_662837\|20 | ···················· | ···················· | ····················· | ···················· | ···················· | ···················· |
| T | 1326 hCoV-19/England/QEUH-AF46B8/2020\|EPI_ISL_662838\|20 | ···················· | ···················· | ····················· | ···················· | ···················· | ···················· |
| T | 1327 hCoV-19/England/QEUH-AEF96C/2020\|EPI_ISL_662839\|20 | ···················· | ···················· | ····················· | ···················· | ···················· | ···················· |
| T | 1328 hCoV-19/England/QEUH-AF666B/2020\|EPI_ISL_662895\|20 | ···················· | ···················· | ····················· | ···················· | ···················· | ···················· |
| T | 1329 hCoV-19/England/QEUH-AF5550/2020\|EPI_ISL_662897\|20 | ···················· | ···················· | ····················· | ···················· | ···················· | ···················· |
| T | 1330 hCoV-19/England/QEUH-AF57E1/2020\|EPI_ISL_662898\|20 | ···················· | ···················· | ····················· | ···················· | ···················· | ···················· |
| T | 1331 hCoV-19/England/QEUH-AF66A7/2020\|EPI_ISL_662899\|20 | ···················· | ···················· | ····················· | ···················· | ···················· | ···················· |
| T | 1332 hCoV-19/England/CAMB-1BAECE/2020\|EPI_ISL_665000\|20 | ···················· | ···················· | ····················· | ···················· | ···················· | ···················· |
| T | 1333 hCoV-19/England/CAMB-1BB11F/2020\|EPI_ISL_665001\|20 | ···················· | ···················· | ····················· | ···················· | ···················· | ···················· |
| T | 1334 hCoV-19/England/CAMB-1BB16A/2020\|EPI_ISL_665002\|20 | ···················· | ·A·················· | ····················· | ···················· | ···················· | ···················· |
| T | 1335 hCoV-19/England/CAMB-1BB188/2020\|EPI_ISL_665003\|20 | ···················· | ···················· | ····················· | ···················· | ···················· | ···················· |
| T | 1336 hCoV-19/England/CAMB-1BB258/2020\|EPI_ISL_665004\|20 | ···················· | ···················· | ····················· | ···················· | ···················· | ···················· |
| T | 1337 hCoV-19/England/CAMB-1BB2EF/2020\|EPI_ISL_665005\|20 | ···················· | ···················· | ····················· | ···················· | ···················· | ···················· |
| T | 1338 hCoV-19/England/CAMB-1BB346/2020\|EPI_ISL_665006\|20 | ···················· | ···················· | ····················· | ···················· | ···················· | ···················· |
| T | 1339 hCoV-19/England/EXET-13AB83/2020\|EPI_ISL_665090\|20 | ···················· | ···················· | ····················· | ···················· | ······T············· | ···················· |
| T | 1340 hCoV-19/England/EXET-13ABB0/2020\|EPI_ISL_665091\|20 | ···················· | ···················· | ····················· | ···················· | ······T············· | ···················· |
| T | 1341 hCoV-19/England/EXET-13AC80/2020\|EPI_ISL_665092\|20 | ···················· | ···················· | ····················· | ···················· | ······T············· | ···················· |
| T | 1342 hCoV-19/England/EXET-13AD32/2020\|EPI_ISL_665093\|20 | ···················· | ···················· | ····················· | ···················· | ······T············· | ···················· |
| T | 1343 hCoV-19/England/EXET-13AD7E/2020\|EPI_ISL_665094\|20 | ···················· | ···················· | ····················· | ···················· | ······T············· | ···················· |
| T | 1344 hCoV-19/England/EXET-13AD8D/2020\|EPI_ISL_665095\|20 | ···················· | ···················· | ····················· | ···················· | ···················· | ···················· |
| T | 1345 hCoV-19/England/EXET-13ADBA/2020\|EPI_ISL_665096\|20 | ···················· | ···················· | ····················· | ···················· | ······T············· | ···················· |
| T | 1346 hCoV-19/England/EXET-13ADC9/2020\|EPI_ISL_665097\|20 | ···················· | ···················· | ····················· | ···················· | ···················· | ···················· |
| T | 1347 hCoV-19/England/EXET-13ADD8/2020\|EPI_ISL_665098\|20 | ···················· | ···················· | ····················· | ···················· | ···················· | ···················· |
| T | 1348 hCoV-19/England/EXET-13AE3F/2020\|EPI_ISL_665099\|20 | ···················· | ···················· | ····················· | ···················· | ···················· | ···················· |
| T | 1349 hCoV-19/Wales/PHWC-48B986/2020\|EPI_ISL_666305\|2020 | ···················· | ···················· | ····················· | ···················· | ···················· | ···················· |
| T | 1350 hCoV-19/Wales/PHWC-48B9A4/2020\|EPI_ISL_666306\|2020 | ···················· | ···················· | ····················· | ···················· | ···················· | ···················· |
| T | 1351 hCoV-19/Wales/PHWC-48B9B3/2020\|EPI_ISL_666307\|2020 | ···················· | ···················· | ····················· | ···················· | ···················· | ···················· |
| T | 1352 hCoV-19/Wales/PHWC-48B9C2/2020\|EPI_ISL_666308\|2020 | ···················· | ···················· | ····················· | ···················· | ···················· | ···················· |
| T | 1353 hCoV-19/Wales/PHWC-48B9D1/2020\|EPI_ISL_666309\|2020 | ···················· | ···················· | ····················· | ···················· | ···················· | ···················· |
| T | 1354 hCoV-19/Wales/PHWC-48B9E0/2020\|EPI_ISL_666310\|2020 | ···················· | ···················· | ····················· | ···················· | ···················· | ···················· |
| T | 1355 hCoV-19/Wales/PHWC-48B9FF/2020\|EPI_ISL_666311\|2020 | ···················· | ···················· | ····················· | ···················· | ···················· | ···················· |
| T | 1356 hCoV-19/Wales/PHWC-48BA0B/2020\|EPI_ISL_666312\|2020 | ···················· | ···················· | ····················· | ···················· | ···················· | ···················· |
| T | 1357 hCoV-19/Wales/PHWC-48BA29/2020\|EPI_ISL_666314\|2020 | ···················· | ···················· | ····················· | ···················· | ···················· | ···················· |
| T | 1358 hCoV-19/Wales/PHWC-48BA47/2020\|EPI_ISL_666315\|2020 | ···················· | ···················· | ····················· | ···················· | ···················· | ···················· |
| T | 1359 hCoV-19/Wales/PHWC-48BA56/2020\|EPI_ISL_666316\|2020 | ···················· | ···················· | ····················· | ···················· | ···················· | ···················· |
| T | 1360 hCoV-19/Wales/PHWC-48BA74/2020\|EPI_ISL_666317\|2020 | ···················· | ···················· | ····················· | ···················· | ···················· | ···················· |

```
T 1361 hCoV-19/Wales/PHWC-48BA83/2020|EPI_ISL_666318|2020 ···················· ···················· ···················· ···················· ···················· ····················
T 1362 hCoV-19/Wales/PHWC-48BAA1/2020|EPI_ISL_666319|2020 ···················· ···················· ···················· ···················· ···················· ····················
T 1363 hCoV-19/Wales/PHWC-48BACF/2020|EPI_ISL_666320|2020 ···················· ···················· ···················· ···················· ···················· ····················
T 1364 hCoV-19/Wales/PHWC-48BADE/2020|EPI_ISL_666321|2020 ···················· ···················· ···················· ···················· ···················· ····················
T 1365 hCoV-19/Wales/PHWC-48BAED/2020|EPI_ISL_666322|2020 ···················· ···················· ···················· ···················· ···················· ····················
T 1366 hCoV-19/Wales/PHWC-48BAFC/2020|EPI_ISL_666323|2020 ···················· ···················· ···················· ···················· ···················· ····················
T 1367 hCoV-19/Wales/PHWC-48BB08/2020|EPI_ISL_666324|2020 ···················· ···················· ···················· ···················· ···················· ····················
T 1368 hCoV-19/Wales/PHWC-48BB17/2020|EPI_ISL_666325|2020 ···················· ···················· ···················· ···················· ···················· ····················
T 1369 hCoV-19/Wales/PHWC-48BB26/2020|EPI_ISL_666326|2020 ···················· ···················· ···················· ···················· ···················· ····················
T 1370 hCoV-19/Wales/PHWC-48BB35/2020|EPI_ISL_666327|2020 ···················· ···················· ···················· ··············A····· ···················· ····················
T 1371 hCoV-19/Wales/PHWC-48BB44/2020|EPI_ISL_666328|2020 ···················· ···················· ···················· ···················· ···················· ····················
T 1372 hCoV-19/Wales/PHWC-48BB53/2020|EPI_ISL_666329|2020 ···················· ···················· ···················· ···················· ···················· ····················
T 1373 hCoV-19/Wales/PHWC-48BB62/2020|EPI_ISL_666330|2020 ···················· ···················· ···················· ···················· ···················· ····················
T 1374 hCoV-19/Wales/PHWC-48BB71/2020|EPI_ISL_666331|2020 ···················· ···················· ···················· ···················· ···················· ····················
T 1375 hCoV-19/Wales/PHWC-48BBCC/2020|EPI_ISL_666332|2020 ···················· ···················· ···················· ···················· ···················· ····················
T 1376 hCoV-19/Wales/PHWC-48BBDB/2020|EPI_ISL_666333|2020 ···················· ···················· ···················· ···················· ···················· ····················
T 1377 hCoV-19/Wales/PHWC-48BBEA/2020|EPI_ISL_666334|2020 ···················· ···················· ···················· ···················· ···················· ····················
T 1378 hCoV-19/Wales/PHWC-48BBF9/2020|EPI_ISL_666335|2020 ···················· ···················· ···················· ···················· ···················· ····················
T 1379 hCoV-19/Wales/PHWC-48BC14/2020|EPI_ISL_666336|2020 ···················· ···················· ···················· ···················· ···················· ····················
T 1380 hCoV-19/Wales/PHWC-48BC23/2020|EPI_ISL_666337|2020 ···················· ···················· ···················· ···················· ···················· ····················
T 1381 hCoV-19/Japan/IC-0207/2020|EPI_ISL_667636|2020-07 ···················· ···················· ···················· ···················· ···················· ····················
T 1382 hCoV-19/Japan/IC-0208/2020|EPI_ISL_667637|2020-10 ···················· ···················· ···················· ···················· ···················· ····················
T 1383 hCoV-19/Japan/IC-0209/2020|EPI_ISL_667638|2020-10 ···················· ···················· ···················· ···················· ···················· ····················
T 1384 hCoV-19/Japan/IC-0210/2020|EPI_ISL_667639|2020-06 ···················· ···················· ···················· ···················· ···················· ····················
T 1385 hCoV-19/Japan/IC-0211/2020|EPI_ISL_667640|2020-06 ···················· ···················· ···················· ···················· ···················· ····················
T 1386 hCoV-19/Japan/IC-0212/2020|EPI_ISL_667641|2020-06 ···················· ···················· ···················· ···················· ···················· ····················
T 1387 hCoV-19/Japan/IC-0213/2020|EPI_ISL_667642|2020-06 ···················· ···················· ···················· ···················· ···················· ····················
T 1388 hCoV-19/Japan/IC-0214/2020|EPI_ISL_667643|2020-07 ···················· ···················· ···················· ···················· ···················· ····················
T 1389 hCoV-19/Japan/IC-0215/2020|EPI_ISL_667644|2020-07 ···················· ···················· ···················· ···················· ···················· ····················
T 1390 hCoV-19/Japan/IC-0216/2020|EPI_ISL_667645|2020-07 ···················· ···················· ···················· ···················· ···················· ····················
T 1391 hCoV-19/Japan/IC-0217/2020|EPI_ISL_667646|2020-07 ···················· ···················· ···················· ···················· ···················· ····················
T 1392 hCoV-19/Japan/IC-0218/2020|EPI_ISL_667647|2020-07 ···················· ···················· ···················· ···················· ···················· ····················
T 1393 hCoV-19/Japan/IC-0219/2020|EPI_ISL_667648|2020-07 ···················· ···················· ···················· ···················· ···················· ····················
T 1394 hCoV-19/Japan/IC-0220/2020|EPI_ISL_667649|2020-07 ···················· ···················· ···················· ···················· ···················· ····················
T 1395 hCoV-19/Japan/IC-0221/2020|EPI_ISL_667650|2020-07 ···················· ···················· ···················· ···················· ···················· ····················
T 1396 hCoV-19/Japan/IC-0222/2020|EPI_ISL_667651|2020-07 ···················· ···················· ···················· ···················· ···················· ····················
T 1397 hCoV-19/Japan/IC-0223/2020|EPI_ISL_667652|2020-07 ···················· ···················· ···················· ···················· ···················· ····················
T 1398 hCoV-19/Japan/IC-0224/2020|EPI_ISL_667653|2020-08 ···················· ···················· ···················· ···················· ···················· ····················
T 1399 hCoV-19/Japan/IC-0225/2020|EPI_ISL_667654|2020-08 ···················· ···················· ···················· ···················· ···················· ····················
T 1400 hCoV-19/Japan/IC-0226/2020|EPI_ISL_667655|2020-08 ···················· ···················· ···················· ···················· ···················· ····················
T 1401 hCoV-19/Japan/IC-0227/2020|EPI_ISL_667656|2020-08 ···················· ···················· ···················· ···················· ···················· ····················
T 1402 hCoV-19/Japan/IC-0228/2020|EPI_ISL_667657|2020-08 ···················· ···················· ···················· ···················· ···················· ····················
T 1403 hCoV-19/Japan/IC-0229/2020|EPI_ISL_667658|2020-08 ···················· ···················· ···················· ···················· ···················· ····················
T 1404 hCoV-19/Japan/IC-0230/2020|EPI_ISL_667659|2020-08 ···················· ···················· ···················· ···················· ···················· ····················
T 1405 hCoV-19/Japan/IC-0231/2020|EPI_ISL_667660|2020-08 ···················· ···················· ···················· ···················· ···················· ····················
T 1406 hCoV-19/Japan/IC-0232/2020|EPI_ISL_667661|2020-08 ···················· ···················· ···················· ···················· ···················· ····················
T 1407 hCoV-19/Japan/IC-0233/2020|EPI_ISL_667662|2020-08 ···················· ···················· ···················· ···················· ···················· ····················
T 1408 hCoV-19/Japan/IC-0234/2020|EPI_ISL_667663|2020-09 ···················· ···················· ···················· ···················· ···················· ····················
T 1409 hCoV-19/Japan/IC-0235/2020|EPI_ISL_667664|2020-09 ···················· ···················· ···················· ···················· ···················· ····················
T 1410 hCoV-19/Japan/IC-0236/2020|EPI_ISL_667665|2020-09 ···················· ···················· ···················· ···················· ···················· ····················
T 1411 hCoV-19/Japan/IC-0237/2020|EPI_ISL_667666|2020-09 ···················· ···················· ···················· ···················· ···················· ····················
T 1412 hCoV-19/Japan/IC-0238/2020|EPI_ISL_667667|2020-10 ···················· ···················· ···················· ···················· ···················· ····················
T 1413 hCoV-19/Japan/IC-0239/2020|EPI_ISL_667668|2020-10 ···················· ···················· ···················· ···················· ···················· ····················
T 1414 hCoV-19/Japan/IC-0271/2020|EPI_ISL_667700|2020-11 ···················· ···········C········ ···················· ···················· ···················· ····················
T 1415 hCoV-19/Japan/IC-0272/2020|EPI_ISL_667701|2020-11 ···················· ···········C········ ···················· ···················· ···················· ····················
T 1416 hCoV-19/Denmark/DCGC-9431/2020|EPI_ISL_668967|2020 ···················· ···················· ···················· ···················· ···················· ····················
T 1417 hCoV-19/Denmark/DCGC-9433/2020|EPI_ISL_668968|2020 ···················· ···················· ···················· ···················· ···················· ····················
T 1418 hCoV-19/Denmark/DCGC-9435/2020|EPI_ISL_668969|2020 ···················· ···················· ···················· ···················· ···················· ····················
T 1419 hCoV-19/Denmark/DCGC-9436/2020|EPI_ISL_668970|2020 ···················· ···················· ···················· ···················· ···················· ····················
T 1420 hCoV-19/Denmark/DCGC-9438/2020|EPI_ISL_668971|2020 ···················· ···················· ···················· ···················· ···················· ····················
T 1421 hCoV-19/Denmark/DCGC-9439/2020|EPI_ISL_668972|2020 ···················· ···················· ···················· ···················· ···················· ····················
T 1422 hCoV-19/Denmark/DCGC-9440/2020|EPI_ISL_668973|2020 ···················· ···················· ···················· ···················· ···················· ····················
T 1423 hCoV-19/Denmark/DCGC-9442/2020|EPI_ISL_668974|2020 ···················· ···················· ···················· ···················· ···················· ····················
T 1424 hCoV-19/Denmark/DCGC-9444/2020|EPI_ISL_668975|2020 ···················· ···················· ···················· ···················· ···················· ····················
T 1425 hCoV-19/Denmark/DCGC-9449/2020|EPI_ISL_668976|2020 ···················· ···················· ···················· ···················· ···················· ····················
T 1426 hCoV-19/Denmark/DCGC-9450/2020|EPI_ISL_668977|2020 ···················· ···················· ···················· ···················· ···················· ····················
T 1427 hCoV-19/Denmark/DCGC-9452/2020|EPI_ISL_668978|2020 ···················· ···················· ···················· ···················· ···················· ····················
T 1428 hCoV-19/Denmark/DCGC-9454/2020|EPI_ISL_668979|2020 ···················· ···················· ···················· ···················· ···················· ····················
T 1429 hCoV-19/Denmark/DCGC-9455/2020|EPI_ISL_668980|2020 ···················· ···················· ···················· ···················· ···················· ····················
T 1430 hCoV-19/Denmark/DCGC-9458/2020|EPI_ISL_668981|2020 ···················· ···················· ···················· ···················· ···················· ····················
T 1431 hCoV-19/Denmark/DCGC-9459/2020|EPI_ISL_668982|2020 ···················· ···················· ···················· ···················· ···················· ····················
T 1432 hCoV-19/Denmark/DCGC-9460/2020|EPI_ISL_668983|2020 ···················· ···················· ···················· ···················· ···················· ····················
T 1433 hCoV-19/Denmark/DCGC-9461/2020|EPI_ISL_668984|2020 ···················· ···················· ···················· ···················· ···················· ····················
T 1434 hCoV-19/Denmark/DCGC-9468/2020|EPI_ISL_668985|2020 ···················· ···················· ···················· ···················· ···················· ····················
T 1435 hCoV-19/Denmark/DCGC-9469/2020|EPI_ISL_668986|2020 ···················· ···················· ···················· ···················· ···················· ····················
T 1436 hCoV-19/Denmark/DCGC-9470/2020|EPI_ISL_668987|2020 ···················· ···················· ···················· ···················· ···················· ····················
T 1437 hCoV-19/Denmark/DCGC-9471/2020|EPI_ISL_668988|2020 ···················· ···················· ···················· ···················· ···················· ····················
T 1438 hCoV-19/Denmark/DCGC-9472/2020|EPI_ISL_668989|2020 ···················· ···················· ···················· ···················· ···················· ····················
T 1439 hCoV-19/Denmark/DCGC-9476/2020|EPI_ISL_668990|2020 ···················· ·A·················· ···················· ···················· ···················· ····················
T 1440 hCoV-19/Denmark/DCGC-9478/2020|EPI_ISL_668991|2020 ···················· ···················· ···················· ···················· ···················· ····················
```

T 1441 hCoV-19/Denmark/DCGC-9481/2020|EPI_ISL_668992|2020 ···················· ···················· ····················· ···················· ···················· ····················
T 1442 hCoV-19/Denmark/DCGC-9483/2020|EPI_ISL_668993|2020 ···················· ···················· ····················· ···················· ···················· ····················
T 1443 hCoV-19/Denmark/DCGC-9484/2020|EPI_ISL_668994|2020 ···················· ···················· ····················· ···················· ···················· ····················
T 1444 hCoV-19/Denmark/DCGC-9485/2020|EPI_ISL_668995|2020 ···················· ·A·················· ····················· ···················· ···················· ····················
T 1445 hCoV-19/Denmark/DCGC-9486/2020|EPI_ISL_668996|2020 ···················· ···················· ····················· ···················· ···················· ····················
T 1446 hCoV-19/Denmark/DCGC-9488/2020|EPI_ISL_668997|2020 ···················· ···················· ····················· ···················· ···················· ····················
T 1447 hCoV-19/Denmark/DCGC-9489/2020|EPI_ISL_668998|2020 ···················· ···················· ····················· ···················· ···················· ····················
T 1448 hCoV-19/Denmark/DCGC-9490/2020|EPI_ISL_668999|2020 ···················· ···················· ····················· ···················· ···················· ····················
T 1450 hCoV-19/USA/CA-CZB-10099/2020|EPI_ISL_672213|2020- ···················· ···················· ····················· ···················· ···················· ····················
T 1451 hCoV-19/USA/CA-CZB-10102/2020|EPI_ISL_672214|2020- ···················· ···················· ····················· ···················· ···················· ····················
T 1452 hCoV-19/USA/CA-CZB-10105/2020|EPI_ISL_672215|2020- ···················· ·A·················· ····················· ···················· ···················· ····················
T 1454 hCoV-19/USA/CA-CZB-10109/2020|EPI_ISL_672217|2020- ···················· ·A·················· ····················· ···················· ···················· ····················
T 1455 hCoV-19/USA/CA-CZB-10110/2020|EPI_ISL_672218|2020- ···················· ···················· ····················· ···················· ···················· ····················
T 1456 hCoV-19/USA/CA-CZB-10112/2020|EPI_ISL_672219|2020- ···················· ···················· ····················· ···················· ···················· ····················
T 1457 hCoV-19/USA/CA-CZB-10117/2020|EPI_ISL_672220|2020- ···················· ···················· ····················· ···················· ···················· ····················
T 1458 hCoV-19/USA/CA-CZB-10120/2020|EPI_ISL_672221|2020- ···················· ···················· ····················· ···················· ···················· ····················
T 1460 hCoV-19/USA/CA-CZB-10128/2020|EPI_ISL_672223|2020- ···················· ···················· ····················· ···················· ···················· ····················
T 1461 hCoV-19/USA/CA-CZB-10130/2020|EPI_ISL_672224|2020- ···················· ···················· ····················· ···················· ···················· ····················
T 1462 hCoV-19/USA/CA-CZB-10133/2020|EPI_ISL_672225|2020- ···················· ···················· ····················· ···················· ···················· ····················
T 1463 hCoV-19/USA/CA-CZB-10134/2020|EPI_ISL_672226|2020- ···················· ···················· ····················· ···················· ···················· ····················
T 1464 hCoV-19/USA/CA-CZB-10138/2020|EPI_ISL_672227|2020- ···················· ·A·················· ····················· ···················· ···················· ····················
T 1465 hCoV-19/USA/CA-CZB-10139/2020|EPI_ISL_672228|2020- ···················· ·A·················· ····················· ···················· ···················· ····················
T 1466 hCoV-19/USA/CA-CZB-10140/2020|EPI_ISL_672229|2020- ···················· ···················· ····················· ···················· ···················· ····················
T 1467 hCoV-19/USA/CA-CZB-10143/2020|EPI_ISL_672230|2020- ···················· ···················· ····················· ···················· ···················· ····················
T 1468 hCoV-19/USA/CA-CZB-10144/2020|EPI_ISL_672231|2020- ···················· ·A·················· ····················· ···················· ···················· ····················
T 1469 hCoV-19/USA/CA-CZB-10149/2020|EPI_ISL_672232|2020- ···················· ···················· ····················· ···················· ···················· ····················
T 1470 hCoV-19/USA/CA-CZB-10150/2020|EPI_ISL_672233|2020- ···················· ·A·················· ····················· ···················· ···················· ····················
T 1471 hCoV-19/USA/CA-CZB-10152/2020|EPI_ISL_672234|2020- ···················· ···················· ····················· ···················· ···················· ····················
T 1473 hCoV-19/USA/CA-CZB-10158/2020|EPI_ISL_672236|2020- ···················· ·A·················· ····················· ···················· ···················· ····················
T 1474 hCoV-19/USA/CA-CZB-10160/2020|EPI_ISL_672237|2020- ···················· ···················· ····················· ···················· ···················· ····················
T 1475 hCoV-19/USA/CA-CZB-10161/2020|EPI_ISL_672238|2020- ···················· ···················· ····················· ···················· ···················· ····················
T 1476 hCoV-19/USA/CA-CZB-10162/2020|EPI_ISL_672239|2020- ···················· ···················· ····················· ···················· ···················· ····················
T 1477 hCoV-19/USA/CA-CZB-10165/2020|EPI_ISL_672240|2020- ···················· ···················· ····················· ···················· ···················· ····················
T 1478 hCoV-19/USA/CA-CZB-10166/2020|EPI_ISL_672241|2020- ···················· ···················· ····················· ···················· ···················· ····················
T 1479 hCoV-19/USA/CA-CZB-10168/2020|EPI_ISL_672242|2020- ···················· ···················· ····················· ···················· ···················· ····················
T 1480 hCoV-19/USA/CA-CZB-10169/2020|EPI_ISL_672243|2020- ···················· ···················· ····················· ···················· ···················· ····················
T 1481 hCoV-19/USA/CA-CZB-10172/2020|EPI_ISL_672244|2020- ···················· ···················· ····················· ···················· ···················· ····················
T 1482 hCoV-19/USA/CA-CZB-10176/2020|EPI_ISL_672245|2020- ···················· ···················· ····················· ···················· ···················· ····················
T 1484 hCoV-19/USA/CA-CZB-10182/2020|EPI_ISL_672247|2020- ···················· ···················· ····················· ···················· ···················· ····················
T 1485 hCoV-19/USA/CA-CZB-10183/2020|EPI_ISL_672248|2020- ···················· ···················· ····················· ···················· ···················· ····················
T 1486 hCoV-19/USA/CA-CZB-10184/2020|EPI_ISL_672249|2020- ···················· ···················· ····················· ···················· ···················· ····················
T 1487 hCoV-19/USA/CA-CZB-10185/2020|EPI_ISL_672250|2020- ···················· ···················· ····················· ···················· ···················· ····················
T 1488 hCoV-19/USA/CA-CZB-10186/2020|EPI_ISL_672251|2020- ···················· ···················· ····················· ···················· ···················· ····················
T 1489 hCoV-19/USA/CA-CZB-10188/2020|EPI_ISL_672252|2020- ···················· ·A·················· ····················· ···················· ···················· ····················
T 1490 hCoV-19/USA/CA-CZB-10191/2020|EPI_ISL_672253|2020- ···················· ···················· ····················· ···················· ···················· ····················
T 1491 hCoV-19/USA/CA-CZB-10193/2020|EPI_ISL_672254|2020- ···················· ···················· ····················· ···················· ···················· ····················
T 1492 hCoV-19/USA/CA-CZB-10194/2020|EPI_ISL_672255|2020- ···················· ···················· ····················· ···················· ···················· ····················
T 1494 hCoV-19/USA/CA-CZB-10199/2020|EPI_ISL_672257|2020- ···················· ···················· ····················· ···················· ···················· ····················
T 1495 hCoV-19/USA/CA-CZB-10202/2020|EPI_ISL_672258|2020- ···················· ···················· ····················· ···················· ···················· ····················
T 1496 hCoV-19/USA/CA-CZB-10211/2020|EPI_ISL_672259|2020- ···················· ···················· ····················· ···················· ···················· ····················
T 1497 hCoV-19/USA/CA-CZB-10212/2020|EPI_ISL_672260|2020- ···················· ···················· ····················· ···················· ··N················· ····················
T 1498 hCoV-19/USA/CA-CZB-10218/2020|EPI_ISL_672261|2020- ···················· ···················· ····················· ···················· ···················· ····················
T 1499 hCoV-19/USA/CA-CZB-10220/2020|EPI_ISL_672262|2020- ···················· ···················· ····················· ···················· ···················· ····················
T 1500 hCoV-19/USA/CA-CZB-10221/2020|EPI_ISL_672263|2020- ···················· ···················· ····················· ···················· ···················· ····················
T 1501 hCoV-19/USA/CA-CZB-10225/2020|EPI_ISL_672264|2020- ···················· ···················· ····················· ···················· ···················· ····················
T 1502 hCoV-19/USA/CA-CZB-10227/2020|EPI_ISL_672265|2020- ···················· ···················· ····················· ···················· ···················· ····················
T 1503 hCoV-19/USA/CA-CZB-10230/2020|EPI_ISL_672266|2020- ···················· ···················· ····················· ···················· ···················· ····················
T 1505 hCoV-19/USA/CA-CZB-10234/2020|EPI_ISL_672268|2020- ···················· ···················· ····················· ···················· ···················· ····················
T 1506 hCoV-19/USA/CA-CZB-10236/2020|EPI_ISL_672269|2020- ···················· ···················· ····················· ···················· ···················· ····················
T 1507 hCoV-19/USA/CA-CZB-10242/2020|EPI_ISL_672270|2020- ···················· ···················· ····················· ···················· ···················· ····················
T 1508 hCoV-19/USA/CA-CZB-10245/2020|EPI_ISL_672271|2020- ···················· ···················· ····················· ···················· ···················· ····················
T 1509 hCoV-19/USA/CA-CZB-10251/2020|EPI_ISL_672272|2020- ···················· ···················· ····················· ···················· ···················· ····················
T 1510 hCoV-19/USA/CA-CZB-10252/2020|EPI_ISL_672273|2020- ···················· ···················· ····················· ···················· ···················· ····················
T 1511 hCoV-19/USA/CA-CZB-10253/2020|EPI_ISL_672274|2020- ···················· ···················· ····················· ···················· ···················· ····················
T 1512 hCoV-19/USA/CA-CZB-11877/2020|EPI_ISL_672275|2020- ···················· ···················· ····················· ···················· ···················· ····················
T 1513 hCoV-19/USA/CA-CZB-11878/2020|EPI_ISL_672276|2020- ···················· ···················· ····················· ···················· ···················· ····················
T 1514 hCoV-19/USA/CA-CZB-11880/2020|EPI_ISL_672277|2020- ···················· ·A·················· ····················· ···················· ···················· ····················
T 1516 hCoV-19/USA/CA-CZB-11882/2020|EPI_ISL_672279|2020- ···················· ·A·················· ····················· ···················· ···················· ····················
T 1517 hCoV-19/USA/CA-CZB-11886/2020|EPI_ISL_672280|2020- ···················· ·A·················· ····················· ···················· ···················· ····················
T 1518 hCoV-19/USA/CA-CZB-11887/2020|EPI_ISL_672281|2020- ···················· ·A·················· ····················· ···················· ···················· ····················
T 1519 hCoV-19/USA/CA-CZB-11890/2020|EPI_ISL_672282|2020- ···················· ···················· ····················· ···················· ···················· ····················
T 1520 hCoV-19/USA/CA-CZB-11894/2020|EPI_ISL_672283|2020- ···················· ·A·················· ····················· ···················· ···················· ····················
T 1521 hCoV-19/USA/CA-CZB-11898/2020|EPI_ISL_672284|2020- ···················· ·A·················· ····················· ···················· ···················· ····················
T 1522 hCoV-19/USA/CA-CZB-11900/2020|EPI_ISL_672285|2020- ···················· ·A·················· ····················· ···················· ···················· ····················
T 1523 hCoV-19/USA/CA-CZB-11901/2020|EPI_ISL_672286|2020- ···················· ·A·················· ····················· ···················· ···················· ····················
T 1524 hCoV-19/USA/CA-CZB-11911/2020|EPI_ISL_672287|2020- ···················· ·A·················· ····················· ···················· ···················· ····················
T 1526 hCoV-19/USA/CA-CZB-11914/2020|EPI_ISL_672289|2020- ···················· ·A·················· ····················· ···················· ···················· ····················
T 1528 hCoV-19/USA/CA-CZB-11919/2020|EPI_ISL_672291|2020- ···················· ·A·················· ····················· ···················· ···················· ····················
T 1529 hCoV-19/USA/CA-CZB-12975/2020|EPI_ISL_672292|2020- ···················· ···················· ····················· ···················· ···················· ····················
T 1530 hCoV-19/USA/CA-CZB-12981/2020|EPI_ISL_672293|2020- ···················· ···················· ····················· ···················· ···················· ····················

| | | | | | | | | |
|---|---|---|---|---|---|---|---|---|
| T | 1531 | hCoV-19/USA/CA-CZB-12983/2020\|EPI_ISL_672294\|2020- | ···················· | ···················· | ····················· | ···················· | ···················· | ···················· |
| T | 1532 | hCoV-19/USA/CA-CZB-12988/2020\|EPI_ISL_672295\|2020- | ···················· | ···················· | ····················· | ···················· | ···················· | ···················· |
| T | 1533 | hCoV-19/USA/CA-CZB-12994/2020\|EPI_ISL_672296\|2020- | ···················· | ···················· | ····················· | ···················· | ···················· | ···················· |
| T | 1534 | hCoV-19/USA/CA-CZB-13000/2020\|EPI_ISL_672297\|2020- | ···················· | ·A·················· | ····················· | ···················· | ···················· | ···············A···· |
| T | 1535 | hCoV-19/USA/CA-CZB-13009/2020\|EPI_ISL_672298\|2020- | ···················· | ···················· | ····················· | ···················· | ···················· | ···················· |
| T | 1536 | hCoV-19/USA/CA-CZB-13013/2020\|EPI_ISL_672299\|2020- | ···················· | ···················· | ····················· | ···················· | ···················· | ···················· |
| T | 1537 | hCoV-19/England/QEUH-AF476A/2020\|EPI_ISL_662800\|20 | ···················· | ···················· | ····················· | ···················· | ···················· | ···················· |
| T | 1538 | hCoV-19/England/QEUH-AF474C/2020\|EPI_ISL_662802\|20 | ···················· | ···················· | ····················· | ···················· | ···················· | ···················· |
| T | 1539 | hCoV-19/England/QEUH-AF282D/2020\|EPI_ISL_662803\|20 | ···················· | ···················· | ····················· | ···················· | ···················· | ···················· |
| T | 1540 | hCoV-19/England/QEUH-AF0FC0/2020\|EPI_ISL_662805\|20 | ···················· | ·A·················· | ····················· | ···················· | ···················· | ···················· |
| T | 1541 | hCoV-19/England/QEUH-AEFF88/2020\|EPI_ISL_662808\|20 | ···················· | ···················· | ····················· | ···················· | ···················· | ···················· |
| T | 1542 | hCoV-19/England/QEUH-AF0072/2020\|EPI_ISL_662809\|20 | ···················· | ···················· | ····················· | ···················· | ···················· | ···················· |
| T | 1543 | hCoV-19/England/CAMB-1BB6E3/2020\|EPI_ISL_665018\|20 | ···················· | ···················· | ····················· | ···················· | ······T············· | ···················· |
| T | 1544 | hCoV-19/England/CAMB-1BB768/2020\|EPI_ISL_665019\|20 | ···················· | ···················· | ····················· | ···················· | ···················· | ···················· |
| T | 1545 | hCoV-19/England/CAMB-1BB777/2020\|EPI_ISL_665020\|20 | ···················· | ···················· | ····················· | ···················· | ···················· | ···················· |
| T | 1546 | hCoV-19/England/CAMB-1BB7D1/2020\|EPI_ISL_665021\|20 | ···················· | ···················· | ····················· | ···················· | ···················· | ···················· |
| T | 1547 | hCoV-19/England/CAMB-1BB838/2020\|EPI_ISL_665022\|20 | ···················· | ···················· | ····················· | ···················· | ···················· | ···················· |
| T | 1548 | hCoV-19/England/CAMC-B22FF1/2020\|EPI_ISL_665023\|20 | ···················· | ···················· | ····················· | ···················· | ···················· | ···················· |
| T | 1549 | hCoV-19/England/CAMC-B2300C/2020\|EPI_ISL_665024\|20 | ···················· | ···················· | ····················· | ···················· | ···················· | ···················· |
| T | 1550 | hCoV-19/England/CAMC-B2302A/2020\|EPI_ISL_665026\|20 | ···················· | ···················· | ····················· | ···················· | ···················· | ···················· |
| T | 1551 | hCoV-19/England/CAMC-B231EB/2020\|EPI_ISL_665029\|20 | ···················· | ···················· | ····················· | ···················· | ···················· | ···················· |
| T | 1552 | hCoV-19/England/CAMC-B231FA/2020\|EPI_ISL_665030\|20 | ···················· | ···················· | ····················· | ···················· | ······T············· | ···················· |
| T | 1553 | hCoV-19/England/CAMC-B23206/2020\|EPI_ISL_665031\|20 | ···················· | ···················· | ····················· | ···················· | ······T············· | ···················· |
| T | 1554 | hCoV-19/England/CAMC-B23321/2020\|EPI_ISL_665032\|20 | ···················· | ···················· | ····················· | ···················· | ···················· | ···················· |
| T | 1555 | hCoV-19/England/CAMC-B2334F/2020\|EPI_ISL_665033\|20 | ···················· | ···················· | ····················· | ···················· | ···················· | ···················· |
| T | 1556 | hCoV-19/England/CAMC-B2337C/2020\|EPI_ISL_665034\|20 | ···················· | ···················· | ····················· | ···················· | ···················· | ···················· |
| T | 1557 | hCoV-19/England/CAMC-B233A9/2020\|EPI_ISL_665035\|20 | ···················· | ···················· | ····················· | ···················· | ···················· | ···················· |
| T | 1558 | hCoV-19/England/CAMC-B2343D/2020\|EPI_ISL_665036\|20 | ···················· | ···················· | ····················· | ···················· | ···················· | ···················· |
| T | 1559 | hCoV-19/England/CAMC-B2344C/2020\|EPI_ISL_665037\|20 | ···················· | ···················· | ····················· | ···················· | ···················· | ···················· |
| T | 1560 | hCoV-19/England/CAMC-B2346A/2020\|EPI_ISL_665038\|20 | ···················· | ···················· | ····················· | ···················· | ···················· | ···················· |
| T | 1561 | hCoV-19/England/CAMC-B23479/2020\|EPI_ISL_665039\|20 | ···················· | ·A·················· | ····················· | ···················· | ···················· | ···A················ |
| T | 1562 | hCoV-19/England/CAMC-B234A6/2020\|EPI_ISL_665040\|20 | ···················· | ···················· | ····················· | ···················· | ···················· | ···················· |
| T | 1563 | hCoV-19/England/CAMC-B23549/2020\|EPI_ISL_665041\|20 | ···················· | ···················· | ····················· | ···················· | ···················· | ···················· |
| T | 1564 | hCoV-19/England/CAMC-B23567/2020\|EPI_ISL_665042\|20 | ···················· | ···················· | ····················· | ···················· | ···················· | ···················· |
| T | 1565 | hCoV-19/England/CAMC-B23585/2020\|EPI_ISL_665043\|20 | ···················· | ·A·······A·········· | ····················· | ···················· | ···················· | ···················· |
| T | 1566 | hCoV-19/England/CAMC-B23594/2020\|EPI_ISL_665044\|20 | ···················· | ···················· | ····················· | ···················· | ···················· | ···················· |
| T | 1567 | hCoV-19/England/CAMC-B235A3/2020\|EPI_ISL_665045\|20 | ···················· | ···················· | ····················· | ···················· | ···················· | ···················· |
| T | 1568 | hCoV-19/England/EXET-138DF8/2020\|EPI_ISL_665046\|20 | ···················· | ···················· | ····················· | ···················· | ···················· | ···················· |
| T | 1575 | hCoV-19/England/EXET-1390C1/2020\|EPI_ISL_665053\|20 | ···················· | ···················· | ····················· | ···················· | ···················· | ···················· |
| T | 1578 | hCoV-19/England/EXET-139173/2020\|EPI_ISL_665056\|20 | ···················· | ···················· | ····················· | ···················· | ······T············· | ···················· |
| T | 1584 | hCoV-19/England/EXET-1392F8/2020\|EPI_ISL_665062\|20 | ···················· | ···················· | ····················· | ···················· | ···················· | ···················· |
| T | 1588 | hCoV-19/England/EXET-13A0C0/2020\|EPI_ISL_665066\|20 | ···················· | ···················· | ····················· | ···················· | ···················· | ···················· |
| T | 1589 | hCoV-19/England/EXET-13A3C7/2020\|EPI_ISL_665067\|20 | ···················· | ···················· | ····················· | ···················· | ······T············· | ···················· |
| T | 1590 | hCoV-19/England/EXET-13A637/2020\|EPI_ISL_665068\|20 | ···················· | ···················· | ····················· | ···················· | ······T············· | ···················· |
| T | 1591 | hCoV-19/England/EXET-13A6CE/2020\|EPI_ISL_665069\|20 | ···················· | ···················· | ····················· | ···················· | ······T············· | ···················· |
| T | 1592 | hCoV-19/England/EXET-13A725/2020\|EPI_ISL_665070\|20 | ···················· | ···················· | ····················· | ···················· | ······T············· | ···················· |
| T | 1593 | hCoV-19/England/EXET-13A743/2020\|EPI_ISL_665071\|20 | ···················· | ···················· | ····················· | ···················· | ······T············· | ···················· |
| T | 1594 | hCoV-19/England/EXET-13A804/2020\|EPI_ISL_665072\|20 | ···················· | ···················· | ····················· | ···················· | ···················· | ···················· |
| T | 1595 | hCoV-19/England/EXET-13A840/2020\|EPI_ISL_665073\|20 | ···················· | ···················· | ····················· | ···················· | ······T············· | ···················· |
| T | 1596 | hCoV-19/England/EXET-13A8B9/2020\|EPI_ISL_665074\|20 | ···················· | ···················· | ····················· | ···················· | ······T············· | ···················· |
| T | 1597 | hCoV-19/England/EXET-13A8D7/2020\|EPI_ISL_665075\|20 | ···················· | ···················· | ····················· | ···················· | ···················· | ···················· |
| T | 1598 | hCoV-19/England/EXET-13A8F5/2020\|EPI_ISL_665076\|20 | ···················· | ···················· | ····················· | ···················· | ······T············· | ···················· |
| T | 1599 | hCoV-19/England/EXET-13A901/2020\|EPI_ISL_665077\|20 | ···················· | ···················· | ····················· | ···················· | ······T············· | ···················· |
| T | 1600 | hCoV-19/England/EXET-13A92F/2020\|EPI_ISL_665078\|20 | ···················· | ···················· | ····················· | ···················· | ······T············· | ···················· |
| T | 1601 | hCoV-19/England/EXET-13A93E/2020\|EPI_ISL_665079\|20 | ···················· | ···················· | ····················· | ···················· | ······T············· | ···················· |
| T | 1602 | hCoV-19/England/EXET-13A97A/2020\|EPI_ISL_665080\|20 | ···················· | ···················· | ····················· | ···················· | ···················· | ···················· |
| T | 1603 | hCoV-19/England/EXET-13AA2C/2020\|EPI_ISL_665081\|20 | ···················· | ···················· | ····················· | ···················· | ······T············· | ···················· |
| T | 1604 | hCoV-19/England/EXET-13AA59/2020\|EPI_ISL_665082\|20 | ···················· | ···················· | ····················· | ···················· | ······T············· | ···················· |
| T | 1605 | hCoV-19/England/EXET-13AAA4/2020\|EPI_ISL_665083\|20 | ···················· | ···················· | ····················· | ···················· | ······T············· | ···················· |
| T | 1606 | hCoV-19/England/EXET-13AAB3/2020\|EPI_ISL_665084\|20 | ···················· | ···················· | ····················· | ···················· | ···················· | ···················· |
| T | 1607 | hCoV-19/England/EXET-13AAC2/2020\|EPI_ISL_665085\|20 | ···················· | ···················· | ····················· | ···················· | ······T············· | ···················· |
| T | 1608 | hCoV-19/England/EXET-13AB1A/2020\|EPI_ISL_665086\|20 | ···················· | ···················· | ····················· | ···················· | ···················· | ···················· |
| T | 1609 | hCoV-19/England/EXET-13AB29/2020\|EPI_ISL_665087\|20 | ···················· | ···················· | ····················· | ···················· | ···················· | ···················· |
| T | 1610 | hCoV-19/England/EXET-13AB47/2020\|EPI_ISL_665088\|20 | ···················· | ···················· | ····················· | ···················· | ······T············· | ···················· |
| T | 1611 | hCoV-19/England/EXET-13AB56/2020\|EPI_ISL_665089\|20 | ···················· | ···················· | ····················· | ···················· | ······T············· | ···················· |
| T | 1612 | hCoV-19/Wales/PHWC-48B91D/2020\|EPI_ISL_666300\|2020 | ···················· | ···················· | ····················· | ···················· | ···················· | ···················· |
| T | 1613 | hCoV-19/Wales/PHWC-48B92C/2020\|EPI_ISL_666301\|2020 | ···················· | ···················· | ····················· | ···················· | ···················· | ···················· |
| T | 1614 | hCoV-19/Wales/PHWC-48B93B/2020\|EPI_ISL_666302\|2020 | ···················· | ···················· | ····················· | ···················· | ···················· | ···················· |
| T | 1615 | hCoV-19/Wales/PHWC-48B959/2020\|EPI_ISL_666303\|2020 | ···················· | ···················· | ····················· | ···················· | ···················· | ···················· |
| T | 1616 | hCoV-19/Wales/PHWC-48B968/2020\|EPI_ISL_666304\|2020 | ···················· | ···················· | ····················· | ···················· | ···················· | ···················· |
| T | 1617 | hCoV-19/Wales/PHWC-48BA1A/2020\|EPI_ISL_666313\|2020 | ···················· | ···················· | ····················· | ···················· | ···················· | ···················· |
| T | 1618 | hCoV-19/Wales/PHWC-48BD20/2020\|EPI_ISL_666349\|2020 | ···················· | ···················· | ····················· | ···················· | ···················· | ···················· |
| T | 1619 | hCoV-19/Wales/PHWC-48BD3F/2020\|EPI_ISL_666350\|2020 | ···················· | ···················· | ····················· | ···················· | ···················· | ···················· |
| T | 1620 | hCoV-19/Wales/PHWC-48BD4E/2020\|EPI_ISL_666351\|2020 | ···················· | ···················· | ····················· | ···················· | ···················· | ···················· |
| T | 1621 | hCoV-19/Wales/PHWC-48BD5D/2020\|EPI_ISL_666352\|2020 | ···················· | ···················· | ····················· | ···················· | ···················· | ···················· |
| T | 1622 | hCoV-19/Wales/PHWC-48BD7B/2020\|EPI_ISL_666353\|2020 | ···················· | ···················· | ····················· | ···················· | ···················· | ···················· |
| T | 1623 | hCoV-19/Wales/PHWC-48BD8A/2020\|EPI_ISL_666354\|2020 | ···················· | ···················· | ····················· | ···················· | ···················· | ···················· |
| T | 1624 | hCoV-19/Wales/PHWC-48BD99/2020\|EPI_ISL_666355\|2020 | ···················· | ···················· | ····················· | ···················· | ···················· | ···················· |
| T | 1625 | hCoV-19/Wales/PHWC-48BDC6/2020\|EPI_ISL_666358\|2020 | ···················· | ···················· | ····················· | ···················· | ···················· | ···················· |
| T | 1626 | hCoV-19/Wales/PHWC-48BE1E/2020\|EPI_ISL_666360\|2020 | ···················· | ···················· | ····················· | ···················· | ···················· | ···················· |

| | | | | | | | | |
|---|---|---|---|---|---|---|---|---|
| T | 1627 hCoV-19/Wales/PHWC-48BE5A/2020\|EPI_ISL_666361\|2020 | ···················· | ···················· | ···················· | ···················· | ···················· | ···················· | |
| T | 1628 hCoV-19/Wales/PHWC-48BE78/2020\|EPI_ISL_666362\|2020 | ···················· | ···················· | ···················· | ···················· | ···················· | ···················· | |
| T | 1629 hCoV-19/Wales/PHWC-48BE96/2020\|EPI_ISL_666363\|2020 | ···················· | ···················· | ···················· | ···················· | ···················· | ···················· | |
| T | 1630 hCoV-19/Wales/PHWC-48BEA5/2020\|EPI_ISL_666364\|2020 | ···················· | ···················· | ···················· | ···················· | ···················· | ···················· | |
| T | 1631 hCoV-19/Wales/PHWC-48BEB4/2020\|EPI_ISL_666365\|2020 | ···················· | ···················· | ···················· | ···················· | ···················· | ···················· | |
| T | 1632 hCoV-19/Wales/PHWC-48BEC3/2020\|EPI_ISL_666366\|2020 | ···················· | ···················· | ···················· | ···················· | ···················· | ···················· | |
| T | 1633 hCoV-19/Wales/PHWC-48BED2/2020\|EPI_ISL_666367\|2020 | ···················· | ···················· | ···················· | ···················· | ···················· | ···················· | |
| T | 1634 hCoV-19/Wales/PHWC-48BEF0/2020\|EPI_ISL_666368\|2020 | ···················· | ···················· | ···················· | ···················· | ···················· | ···················· | |
| T | 1635 hCoV-19/Wales/PHWC-48BF0C/2020\|EPI_ISL_666369\|2020 | ···················· | ···················· | ···················· | ···················· | ···················· | ···················· | |
| T | 1636 hCoV-19/Wales/PHWC-48BF1B/2020\|EPI_ISL_666370\|2020 | ···················· | ···················· | ···················· | ···················· | ···················· | ···················· | |
| T | 1637 hCoV-19/Wales/PHWC-48BF39/2020\|EPI_ISL_666371\|2020 | ···················· | ···················· | ···················· | ···················· | ···················· | ···················· | |
| T | 1638 hCoV-19/Wales/PHWC-48BF48/2020\|EPI_ISL_666372\|2020 | ···················· | ···················· | ···················· | ···················· | ···················· | ···················· | |
| T | 1639 hCoV-19/Wales/PHWC-48BF57/2020\|EPI_ISL_666373\|2020 | ···················· | ···················· | ···················· | ···················· | ···················· | ···················· | |
| T | 1640 hCoV-19/Wales/PHWC-48BF84/2020\|EPI_ISL_666374\|2020 | ···················· | ···················· | ···················· | ···················· | ···················· | ···················· | |
| T | 1641 hCoV-19/Wales/PHWC-48BFA2/2020\|EPI_ISL_666375\|2020 | ···················· | ···················· | ···················· | ···················· | ···················· | ···················· | |
| T | 1642 hCoV-19/Wales/PHWC-48BFC0/2020\|EPI_ISL_666376\|2020 | ···················· | ···················· | ···················· | ···················· | ···················· | ···················· | |
| T | 1643 hCoV-19/Wales/PHWC-48BFDF/2020\|EPI_ISL_666377\|2020 | ···················· | ···················· | ···················· | ···················· | ···················· | ···················· | |
| T | 1644 hCoV-19/Wales/PHWC-48BFEE/2020\|EPI_ISL_666378\|2020 | ···················· | ···················· | ···················· | ···················· | ···················· | ···················· | |
| T | 1645 hCoV-19/Wales/PHWC-48BFFD/2020\|EPI_ISL_666379\|2020 | ···················· | ···················· | ···················· | ···················· | ···················· | ···················· | |
| T | 1646 hCoV-19/Wales/PHWC-48C017/2020\|EPI_ISL_666380\|2020 | ···················· | ···················· | ···················· | ···················· | ···················· | ···················· | |
| T | 1647 hCoV-19/Wales/PHWC-48C026/2020\|EPI_ISL_666381\|2020 | ···················· | ···················· | ···················· | ···················· | ···················· | ···················· | |
| T | 1648 hCoV-19/Wales/PHWC-48C035/2020\|EPI_ISL_666382\|2020 | ···················· | ···················· | ···················· | ···················· | ···················· | ···················· | |
| T | 1649 hCoV-19/Wales/PHWC-48C044/2020\|EPI_ISL_666383\|2020 | ···················· | ···················· | ···················· | ···················· | ···················· | ···················· | |
| T | 1650 hCoV-19/Wales/PHWC-48C053/2020\|EPI_ISL_666384\|2020 | ···················· | ···················· | ···················· | ···················· | NNN················· | ···················· | 1 |
| T | 1651 hCoV-19/Wales/PHWC-48C062/2020\|EPI_ISL_666385\|2020 | ···················· | ···················· | ···················· | ···················· | ···················· | ···················· | |
| T | 1652 hCoV-19/Wales/PHWC-48C071/2020\|EPI_ISL_666386\|2020 | ···················· | ···················· | ···················· | ···················· | ···················· | ···················· | |
| T | 1653 hCoV-19/Wales/PHWC-48C080/2020\|EPI_ISL_666387\|2020 | ···················· | ···················· | ···················· | ···················· | NNN················· | ···················· | 1 |
| T | 1654 hCoV-19/Wales/PHWC-48C09F/2020\|EPI_ISL_666388\|2020 | ···················· | ···················· | ···················· | ···················· | ···················· | ···················· | |
| T | 1655 hCoV-19/Wales/PHWC-48C0AE/2020\|EPI_ISL_666389\|2020 | ···················· | ···················· | ···················· | ···················· | ···················· | ···················· | |
| T | 1656 hCoV-19/Wales/PHWC-48C0BD/2020\|EPI_ISL_666390\|2020 | ···················· | ···················· | ···················· | ···················· | ···················· | ···················· | |
| T | 1657 hCoV-19/Wales/PHWC-48C0DB/2020\|EPI_ISL_666391\|2020 | ···················· | ···················· | ···················· | ···················· | ···················· | ···················· | |
| T | 1658 hCoV-19/Wales/PHWC-48C0EA/2020\|EPI_ISL_666392\|2020 | ···················· | ···················· | ···················· | ···················· | ···················· | ···················· | |
| T | 1659 hCoV-19/Wales/PHWC-48C0F9/2020\|EPI_ISL_666393\|2020 | ···················· | ···················· | ···················· | ···················· | ···················· | ···················· | |
| T | 1660 hCoV-19/Wales/PHWC-48C105/2020\|EPI_ISL_666394\|2020 | ···················· | ·A·················· | ···················· | ···················· | ···················· | ··A················· | |
| T | 1661 hCoV-19/Wales/PHWC-48C114/2020\|EPI_ISL_666395\|2020 | ···················· | ···················· | ···················· | ···················· | ···················· | ···················· | |
| T | 1662 hCoV-19/Wales/PHWC-48C123/2020\|EPI_ISL_666396\|2020 | ···················· | ···················· | ···················· | ···················· | ···················· | ···················· | |
| T | 1663 hCoV-19/Wales/PHWC-48C132/2020\|EPI_ISL_666397\|2020 | ···················· | ···················· | ···················· | ···················· | ···················· | ···················· | |
| T | 1665 hCoV-19/Wales/PHWC-48C150/2020\|EPI_ISL_666399\|2020 | ···················· | ···················· | ···················· | ···················· | ···················· | ···················· | |
| T | 1666 hCoV-19/Japan/IC-0171/2020\|EPI_ISL_667600\|2020-09 | ···················· | ···················· | ···················· | ···················· | ···················· | ···················· | |
| T | 1667 hCoV-19/Japan/IC-0172/2020\|EPI_ISL_667601\|2020-09 | ···················· | ···················· | ···················· | ···················· | ···················· | ···················· | |
| T | 1668 hCoV-19/Japan/IC-0173/2020\|EPI_ISL_667602\|2020-09 | ···················· | ·A·················· | ···················· | ···················· | ···················· | ··A················· | |
| T | 1669 hCoV-19/Japan/IC-0174/2020\|EPI_ISL_667603\|2020-09 | ···················· | ···················· | ···················· | ···················· | ···················· | ···················· | |
| T | 1670 hCoV-19/Japan/IC-0175/2020\|EPI_ISL_667604\|2020-09 | ···················· | ···················· | ···················· | ···················· | ···················· | ···················· | |
| T | 1671 hCoV-19/Japan/IC-0176/2020\|EPI_ISL_667605\|2020-09 | ·····G·············· | ···················· | ···················· | ···················· | ···················· | ···················· | |
| T | 1672 hCoV-19/Japan/IC-0177/2020\|EPI_ISL_667606\|2020-09 | ···················· | ···················· | ···················· | ···················· | ···················· | ···················· | |
| T | 1673 hCoV-19/Japan/IC-0178/2020\|EPI_ISL_667607\|2020-09 | ···················· | ·A·················· | ···················· | ···················· | ···················· | ···················· | |
| T | 1674 hCoV-19/Japan/IC-0179/2020\|EPI_ISL_667608\|2020-09 | ···················· | ···················· | ···················· | ···················· | ···················· | ···················· | |
| T | 1675 hCoV-19/Japan/IC-0180/2020\|EPI_ISL_667609\|2020-09 | ···················· | ·A·················· | ···················· | ···················· | ···················· | ··A················· | |
| T | 1676 hCoV-19/Japan/IC-0181/2020\|EPI_ISL_667610\|2020-09 | ···················· | ···················· | ···················· | ···················· | ···················· | ···················· | |
| T | 1677 hCoV-19/Japan/IC-0182/2020\|EPI_ISL_667611\|2020-10 | ···················· | ···················· | ···················· | ···················· | ···················· | ···················· | |
| T | 1678 hCoV-19/Japan/IC-0183/2020\|EPI_ISL_667612\|2020-10 | ···················· | ···················· | ···················· | ···················· | ···················· | ···················· | |
| T | 1679 hCoV-19/Japan/IC-0184/2020\|EPI_ISL_667613\|2020-10 | ···················· | ·A·················· | ···················· | ···················· | ···················· | ··A················· | |
| T | 1680 hCoV-19/Japan/IC-0185/2020\|EPI_ISL_667614\|2020-10 | ···················· | ·A·················· | ···················· | ···················· | ···················· | ··A················· | |
| T | 1681 hCoV-19/Japan/IC-0186/2020\|EPI_ISL_667615\|2020-10 | ···················· | ···················· | ···················· | ···················· | ···················· | ···················· | |
| T | 1682 hCoV-19/Japan/IC-0187/2020\|EPI_ISL_667616\|2020-10 | ···················· | ···················· | ···················· | ···················· | ···················· | ···················· | |
| T | 1683 hCoV-19/Japan/IC-0188/2020\|EPI_ISL_667617\|2020-10 | ···················· | ·A·················· | ···················· | ···················· | ···················· | ···················· | |
| T | 1684 hCoV-19/Japan/IC-0190/2020\|EPI_ISL_667619\|2020-10 | ···················· | ···················· | ···················· | ···················· | ···················· | ···················· | |
| T | 1685 hCoV-19/Japan/IC-0191/2020\|EPI_ISL_667620\|2020-10 | ···················· | ···················· | ···················· | ···················· | ···················· | ···················· | |
| T | 1686 hCoV-19/Japan/IC-0193/2020\|EPI_ISL_667622\|2020-06 | ···················· | ···················· | ···················· | ···················· | ···················· | ···················· | |
| T | 1687 hCoV-19/Japan/IC-0194/2020\|EPI_ISL_667623\|2020-07 | ···················· | ···A················ | ···················· | ···················· | ···················· | ···················· | |
| T | 1688 hCoV-19/Japan/IC-0195/2020\|EPI_ISL_667624\|2020-07 | ···················· | ·A·················· | ···················· | ···················· | ···················· | ···················· | |
| T | 1689 hCoV-19/Japan/IC-0196/2020\|EPI_ISL_667625\|2020-07 | ···················· | ·A·················· | ···················· | ···················· | ···················· | ···················· | |
| T | 1690 hCoV-19/Japan/IC-0197/2020\|EPI_ISL_667626\|2020-07 | ···················· | ···················· | ···················· | ···················· | ···················· | ···················· | |
| T | 1691 hCoV-19/Japan/IC-0198/2020\|EPI_ISL_667627\|2020-07 | ···················· | ·A·················· | ···················· | ···················· | ···················· | ···················· | |
| T | 1692 hCoV-19/Japan/IC-0199/2020\|EPI_ISL_667628\|2020-07 | ···················· | ···················· | ···················· | ···················· | ···················· | ···················· | |
| T | 1693 hCoV-19/Japan/IC-0200/2020\|EPI_ISL_667629\|2020-07 | ···················· | ···················· | ···················· | ···················· | ···················· | ···················· | |
| T | 1694 hCoV-19/Japan/IC-0201/2020\|EPI_ISL_667630\|2020-07 | ···················· | ···A················ | ···················· | ···················· | ···················· | ···················· | |
| T | 1695 hCoV-19/Japan/IC-0202/2020\|EPI_ISL_667631\|2020-07 | ···················· | ···················· | ···················· | ···················· | ···················· | ···················· | |
| T | 1696 hCoV-19/Japan/IC-0203/2020\|EPI_ISL_667632\|2020-07 | ···················· | ···················· | ···················· | ···················· | ···················· | ···················· | |
| T | 1697 hCoV-19/Japan/IC-0204/2020\|EPI_ISL_667633\|2020-07 | ···················· | ···················· | ···················· | ···················· | ···················· | ···················· | |
| T | 1698 hCoV-19/Japan/IC-0206/2020\|EPI_ISL_667635\|2020-07 | ···················· | ···················· | ···················· | ···················· | ···················· | ···················· | |
| T | 1699 hCoV-19/Japan/IC-0251/2020\|EPI_ISL_667680\|2020-10 | ···················· | ···················· | ···················· | ···················· | ···················· | ···················· | |
| T | 1700 hCoV-19/Japan/IC-0252/2020\|EPI_ISL_667681\|2020-10 | ···················· | ·A·················· | ···················· | ···················· | ···················· | ··A················· | |
| T | 1701 hCoV-19/Japan/IC-0253/2020\|EPI_ISL_667682\|2020-10 | ···················· | ···················· | ···················· | ···················· | ···················· | ···················· | |
| T | 1702 hCoV-19/Japan/IC-0254/2020\|EPI_ISL_667683\|2020-10 | ···················· | ···················· | ···················· | ···················· | ···················· | ···················· | |
| T | 1703 hCoV-19/Japan/IC-0255/2020\|EPI_ISL_667684\|2020-10 | ···················· | ···················· | ···················· | ···················· | ···················· | ···················· | |
| T | 1704 hCoV-19/Japan/IC-0256/2020\|EPI_ISL_667685\|2020-10 | ···················· | ···················· | ···················· | ···················· | ···················· | ···················· | |
| T | 1705 hCoV-19/Japan/IC-0257/2020\|EPI_ISL_667686\|2020-10 | ···················· | ···················· | ···················· | ···················· | ···················· | ···················· | |
| T | 1706 hCoV-19/Japan/IC-0260/2020\|EPI_ISL_667689\|2020-10 | ···················· | ···················· | ···················· | ···················· | ···················· | ···················· | |
| T | 1707 hCoV-19/Japan/IC-0261/2020\|EPI_ISL_667690\|2020-10 | ···················· | ·A·················· | ···················· | ···················· | ···················· | ···················· | |

T 1708 hCoV-19/Japan/IC-0262/2020|EPI_ISL_667691|2020-10 ···················· ·A·················· ···················· ···················· ···················· ···A················
T 1709 hCoV-19/Japan/IC-0263/2020|EPI_ISL_667692|2020-10 ···················· ···················· ···················· ···················· ···················· ····················
T 1710 hCoV-19/Japan/IC-0264/2020|EPI_ISL_667693|2020-10 ···················· ·A·················· ···················· ···················· ···················· ···A················
T 1711 hCoV-19/Japan/IC-0265/2020|EPI_ISL_667694|2020-11 ···················· ············C······· ···················· ···················· ···················· ····················
T 1712 hCoV-19/Japan/IC-0266/2020|EPI_ISL_667695|2020-11 ···················· ············C······· ···················· ···················· ···················· ····················
T 1713 hCoV-19/Japan/IC-0267/2020|EPI_ISL_667696|2020-11 ···················· ············C······· ···················· ···················· ···················· ····················
T 1714 hCoV-19/Japan/IC-0268/2020|EPI_ISL_667697|2020-11 ···················· ············C······· ···················· ···················· ···················· ····················
T 1715 hCoV-19/Japan/IC-0269/2020|EPI_ISL_667698|2020-11 ···················· ············C······· ···················· ···················· ···················· ····················
T 1716 hCoV-19/Japan/IC-0270/2020|EPI_ISL_667699|2020-11 ···················· ············C······· ···················· ···················· ···················· ····················
T 1717 hCoV-19/Denmark/DCGC-9333/2020|EPI_ISL_668912|2020 ···················· ···················· ···················· ···················· ···················· ····················
T 1718 hCoV-19/Denmark/DCGC-9334/2020|EPI_ISL_668913|2020 ···················· ···················· ···················· ···················· ···················· ····················
T 1719 hCoV-19/Denmark/DCGC-9337/2020|EPI_ISL_668914|2020 ···················· ···················· ···················· ···················· ···················· ····················
T 1720 hCoV-19/Denmark/DCGC-9338/2020|EPI_ISL_668915|2020 ···················· ···················· ···················· ···················· ···················· ····················
T 1721 hCoV-19/Denmark/DCGC-9339/2020|EPI_ISL_668916|2020 ···················· ···················· ···················· ···················· ···················· ····················
T 1722 hCoV-19/Denmark/DCGC-9340/2020|EPI_ISL_668917|2020 ···················· ···················· ···················· ···················· ···················· ····················
T 1723 hCoV-19/Denmark/DCGC-9341/2020|EPI_ISL_668918|2020 ···················· ···················· ···················· ···················· ···················· ····················
T 1724 hCoV-19/Denmark/DCGC-9343/2020|EPI_ISL_668919|2020 ···················· ···················· ···················· ···················· ···················· ····················
T 1725 hCoV-19/Denmark/DCGC-9344/2020|EPI_ISL_668920|2020 ···················· ···················· ···················· ···················· ·······T············ ····················
T 1726 hCoV-19/Denmark/DCGC-9346/2020|EPI_ISL_668921|2020 ···················· ···················· ···················· ···················· ···················· ····················
T 1727 hCoV-19/Denmark/DCGC-9349/2020|EPI_ISL_668922|2020 ···················· ···················· ···················· ···················· ···················· ····················
T 1728 hCoV-19/Denmark/DCGC-9352/2020|EPI_ISL_668923|2020 ···················· ···················· ···················· ···················· ···················· ····················
T 1729 hCoV-19/Denmark/DCGC-9353/2020|EPI_ISL_668924|2020 ···················· ·A·················· ···················· ···················· ···················· ···A················
T 1730 hCoV-19/Denmark/DCGC-9355/2020|EPI_ISL_668925|2020 ···················· ···················· ···················· ···················· ···················· ····················
T 1731 hCoV-19/Denmark/DCGC-9357/2020|EPI_ISL_668926|2020 ···················· ···················· ···················· ···················· ···················· ····················
T 1732 hCoV-19/Denmark/DCGC-9360/2020|EPI_ISL_668927|2020 ···················· ···················· ···················· ···················· ···················· ····················
T 1733 hCoV-19/Denmark/DCGC-9361/2020|EPI_ISL_668928|2020 ···················· ···················· ···················· ···················· ···················· ····················
T 1734 hCoV-19/Denmark/DCGC-9362/2020|EPI_ISL_668929|2020 ···················· ···················· ·········A·········· ···················· ···················· ····················
T 1735 hCoV-19/Denmark/DCGC-9364/2020|EPI_ISL_668930|2020 ···················· ···················· ···················· ···················· ···················· ····················
T 1736 hCoV-19/Denmark/DCGC-9367/2020|EPI_ISL_668931|2020 ···················· ···················· ···················· ···················· ···················· ····················
T 1737 hCoV-19/Denmark/DCGC-9368/2020|EPI_ISL_668932|2020 ···················· ···················· ···················· ···················· ···················· ····················
T 1738 hCoV-19/Denmark/DCGC-9369/2020|EPI_ISL_668933|2020 ···················· ···················· ···················· ···················· ···················· ····················
T 1739 hCoV-19/Denmark/DCGC-9370/2020|EPI_ISL_668934|2020 ···················· ···················· ···················· ···················· ···················· ····················
T 1740 hCoV-19/Denmark/DCGC-9371/2020|EPI_ISL_668935|2020 ···················· ···················· ···················· ···················· ···················· ····················
T 1741 hCoV-19/Denmark/DCGC-9373/2020|EPI_ISL_668936|2020 ···················· ···················· ···················· ···················· ···················· ····················
T 1742 hCoV-19/Denmark/DCGC-9374/2020|EPI_ISL_668937|2020 ···················· ···················· ···················· ···················· ···················· ····················
T 1743 hCoV-19/Denmark/DCGC-9375/2020|EPI_ISL_668938|2020 ···················· ···················· ···················· ···················· ···················· ····················
T 1744 hCoV-19/Denmark/DCGC-9376/2020|EPI_ISL_668939|2020 ···················· ···················· ···················· ···················· ···················· ····················
T 1745 hCoV-19/Denmark/DCGC-9378/2020|EPI_ISL_668940|2020 ···················· ···················· ···················· ···················· ···················· ····················
T 1746 hCoV-19/Denmark/DCGC-9381/2020|EPI_ISL_668941|2020 ···················· ···················· ···················· ···················· ···················· ····················
T 1747 hCoV-19/Denmark/DCGC-9383/2020|EPI_ISL_668942|2020 ···················· ···················· ···················· ···················· ···················· ····················
T 1748 hCoV-19/Denmark/DCGC-9384/2020|EPI_ISL_668943|2020 ···················· ···················· ···················· ···················· ···················· ····················
T 1749 hCoV-19/Denmark/DCGC-9386/2020|EPI_ISL_668944|2020 ···················· ···················· ···················· ···················· ···················· ····················
T 1750 hCoV-19/Denmark/DCGC-9387/2020|EPI_ISL_668945|2020 ···················· ···················· ···················· ···················· ···················· ····················
T 1751 hCoV-19/Denmark/DCGC-9390/2020|EPI_ISL_668946|2020 ···················· ···················· ···················· ···················· ···················· ····················
T 1752 hCoV-19/Denmark/DCGC-9394/2020|EPI_ISL_668947|2020 ···················· ···················· ···················· ···················· ···················· ····················
T 1753 hCoV-19/Denmark/DCGC-9395/2020|EPI_ISL_668948|2020 ···················· ···················· ···················· ···················· ···················· ····················
T 1754 hCoV-19/Denmark/DCGC-9397/2020|EPI_ISL_668949|2020 ···················· ···················· ···················· ···················· ···················· ····················
T 1755 hCoV-19/Denmark/DCGC-9398/2020|EPI_ISL_668950|2020 ···················· ···················· ···················· ···················· ···················· ····················
T 1756 hCoV-19/Denmark/DCGC-9399/2020|EPI_ISL_668951|2020 ···················· ···················· ···················· ···················· ···················· ····················
T 1757 hCoV-19/Denmark/DCGC-9400/2020|EPI_ISL_668952|2020 ···················· ···················· ···················· ···················· ···················· ····················
T 1758 hCoV-19/Denmark/DCGC-9403/2020|EPI_ISL_668953|2020 ···················· ···················· ···················· ···················· ···················· ····················
T 1759 hCoV-19/Denmark/DCGC-9404/2020|EPI_ISL_668954|2020 ···················· ···················· ···················· ···················· ···················· ····················
T 1760 hCoV-19/Denmark/DCGC-9406/2020|EPI_ISL_668955|2020 ···················· ···················· ···················· ···················· ···················· ····················
T 1761 hCoV-19/Denmark/DCGC-9407/2020|EPI_ISL_668956|2020 ···················· ···················· ···················· ···················· ···················· ····················
T 1762 hCoV-19/Denmark/DCGC-9408/2020|EPI_ISL_668957|2020 ···················· ·A·················· ···················· ···················· ···················· ····················
T 1763 hCoV-19/Denmark/DCGC-9412/2020|EPI_ISL_668958|2020 ···················· ···················· ···················· ···················· ···················· ····················
T 1764 hCoV-19/Denmark/DCGC-9415/2020|EPI_ISL_668959|2020 ···················· ·A·················· ···················· ···················· ···················· ···A················
T 1765 hCoV-19/Denmark/DCGC-9417/2020|EPI_ISL_668960|2020 ···················· ···················· ···················· ···················· ···················· ····················
T 1766 hCoV-19/Denmark/DCGC-9419/2020|EPI_ISL_668961|2020 ···················· ···················· ···················· ···················· ···················· ····················
T 1767 hCoV-19/Denmark/DCGC-9420/2020|EPI_ISL_668962|2020 ···················· ···················· ···················· ···················· ···················· ····················
T 1768 hCoV-19/Denmark/DCGC-9421/2020|EPI_ISL_668963|2020 ···················· ···················· ···················· ···················· ···················· ····················
T 1769 hCoV-19/Denmark/DCGC-9423/2020|EPI_ISL_668964|2020 ···················· ···················· ···················· ···················· ···················· ····················
T 1770 hCoV-19/Denmark/DCGC-9427/2020|EPI_ISL_668965|2020 ···················· ···················· ···················· ···················· ···················· ····················
T 1771 hCoV-19/Denmark/DCGC-9429/2020|EPI_ISL_668966|2020 ···················· ···················· ···················· ···················· ···················· ····················
T 1773 hCoV-19/USA/CA-CZB-9794/2020|EPI_ISL_672180|2020-0 ···················· ···················· ···················· ···················· ···················· ····················
T 1774 hCoV-19/USA/CA-CZB-9798/2020|EPI_ISL_672181|2020-0 ···················· ···················· ···················· ···················· ···················· ····················
T 1775 hCoV-19/USA/CA-CZB-9803/2020|EPI_ISL_672182|2020-0 ···················· ···················· ···················· ···················· ···················· ····················
T 1776 hCoV-19/USA/CA-CZB-9998/2020|EPI_ISL_672183|2020-0 ···················· ·A·················· ···················· ···················· ···················· ····················
T 1777 hCoV-19/USA/CA-CZB-10001/2020|EPI_ISL_672184|2020- ···················· ···················· ···················· ···················· ···················· ····················
T 1778 hCoV-19/USA/CA-CZB-10006/2020|EPI_ISL_672185|2020- ···················· ·A·················· ···················· ···················· ···················· ····················
T 1779 hCoV-19/USA/CA-CZB-10007/2020|EPI_ISL_672186|2020- ···················· ··G················· ···················· ···················· ···················· ····················
T 1780 hCoV-19/USA/CA-CZB-10009/2020|EPI_ISL_672187|2020- ···················· ·A·················· ···················· ···················· ···················· ····················
T 1781 hCoV-19/USA/CA-CZB-10010/2020|EPI_ISL_672188|2020- ···················· ··G················· ···················· ···················· ···················· ····················
T 1782 hCoV-19/USA/CA-CZB-10013/2020|EPI_ISL_672189|2020- ···················· ·A·················· ···················· ···················· ···················· ····················
T 1783 hCoV-19/USA/CA-CZB-10014/2020|EPI_ISL_672190|2020- ···················· ···················· ···················· ···················· ···················· ····················
T 1784 hCoV-19/USA/CA-CZB-10015/2020|EPI_ISL_672191|2020- ···················· ···················· ···················· ···················· ···················· ····················
T 1785 hCoV-19/USA/CA-CZB-10025/2020|EPI_ISL_672192|2020- ···················· ·A·················· ···················· ···················· ···················· ····················
T 1786 hCoV-19/USA/CA-CZB-10028/2020|EPI_ISL_672193|2020- ···················· ···················· ···················· ···················· ···················· ····················
T 1787 hCoV-19/USA/CA-CZB-10029/2020|EPI_ISL_672194|2020- ···················· ···················· ···················· ···················· ···················· ····················
T 1788 hCoV-19/USA/CA-CZB-10036/2020|EPI_ISL_672195|2020- ···················· ···················· ···················· ···················· ···················· ····················

| | | | | | | | | |
|---|---|---|---|---|---|---|---|---|
| T | 1789 | hCoV-19/USA/CA-CZB-10039/2020\|EPI_ISL_672196\|2020- | ···················· | ·A·················· | ···················· | ···················· | ···················· | ···················· |
| T | 1790 | hCoV-19/USA/CA-CZB-10040/2020\|EPI_ISL_672197\|2020- | ···················· | ·A·················· | ···················· | ···················· | ···················· | ···················· |
| T | 1791 | hCoV-19/USA/CA-CZB-10042/2020\|EPI_ISL_672198\|2020- | ···················· | ···················· | ···················· | ···················· | ···················· | ···················· |
| T | 1792 | hCoV-19/USA/CA-CZB-10044/2020\|EPI_ISL_672199\|2020- | ···················· | ···················· | ···················· | ···················· | ···················· | ···················· |
| T | 1793 | hCoV-19/England/QEUH-B68997/2020\|EPI_ISL_661400\|20 | ···················· | ·A·················· | ···················· | ···················· | ···················· | ···················· |
| T | 1794 | hCoV-19/England/QEUH-B6829C/2020\|EPI_ISL_661401\|20 | ···················· | ···················· | ···················· | ···················· | ···················· | ···················· |
| T | 1795 | hCoV-19/England/QEUH-B67A59/2020\|EPI_ISL_661403\|20 | ···················· | ···················· | ···················· | ···················· | ···················· | ···················· |
| T | 1796 | hCoV-19/England/QEUH-B679F2/2020\|EPI_ISL_661404\|20 | ···················· | ···················· | ···················· | ···················· | ···················· | ···················· |
| T | 1797 | hCoV-19/England/QEUH-B69A75/2020\|EPI_ISL_661405\|20 | ···················· | ···················· | ···················· | ···················· | ···················· | ···················· |
| T | 1798 | hCoV-19/England/QEUH-B69996/2020\|EPI_ISL_661406\|20 | ···················· | ···················· | ···················· | ···················· | ···················· | ···················· |
| T | 1799 | hCoV-19/England/QEUH-B67C35/2020\|EPI_ISL_661407\|20 | ···················· | ···················· | ···················· | ···················· | ···················· | ···················· |
| T | 1800 | hCoV-19/England/QEUH-B6894C/2020\|EPI_ISL_661408\|20 | ···················· | ···················· | ···················· | ···················· | ···················· | ···················· |
| T | 1801 | hCoV-19/England/QEUH-B679E3/2020\|EPI_ISL_661409\|20 | ···················· | ···················· | ···················· | ···················· | ···················· | ···················· |
| T | 1802 | hCoV-19/England/QEUH-B68A2B/2020\|EPI_ISL_661410\|20 | ···················· | ···················· | ···················· | ···················· | ···················· | ···················· |
| T | 1803 | hCoV-19/England/QEUH-B689F1/2020\|EPI_ISL_661411\|20 | ···················· | ···················· | ···················· | ···················· | ···················· | ···················· |
| T | 1804 | hCoV-19/England/QEUH-B693D4/2020\|EPI_ISL_661412\|20 | ···················· | ···················· | ···················· | ···················· | ···················· | ···················· |
| T | 1805 | hCoV-19/England/QEUH-B69899/2020\|EPI_ISL_661413\|20 | ···················· | ···················· | ···················· | ···················· | ···················· | ···················· |
| T | 1806 | hCoV-19/England/QEUH-B67E4E/2020\|EPI_ISL_661415\|20 | ···················· | ···················· | ···················· | ···················· | ···················· | ···················· |
| T | 1807 | hCoV-19/England/QEUH-B676CE/2020\|EPI_ISL_661416\|20 | ···················· | ···················· | ···················· | ···················· | ···················· | ···················· |
| T | 1808 | hCoV-19/England/QEUH-B67F0F/2020\|EPI_ISL_661417\|20 | ···················· | ···················· | ···················· | ···················· | ···················· | ···················· |
| T | 1809 | hCoV-19/England/QEUH-B6844B/2020\|EPI_ISL_661418\|20 | ···················· | ···················· | ···················· | ···················· | ······T············· | ···················· |
| T | 1810 | hCoV-19/England/QEUH-B66243/2020\|EPI_ISL_661419\|20 | ···················· | ···················· | ···················· | ···················· | ···················· | ···················· |
| T | 1811 | hCoV-19/England/QEUH-B665FF/2020\|EPI_ISL_661420\|20 | ···················· | ·········A·········· | ···················· | ···················· | ······T············· | ···················· |
| T | 1812 | hCoV-19/England/QEUH-B6C508/2020\|EPI_ISL_661421\|20 | ···················· | ···················· | ···················· | ···················· | ···················· | ···················· |
| T | 1813 | hCoV-19/England/QEUH-B68214/2020\|EPI_ISL_661422\|20 | ···················· | ···················· | ···················· | ···················· | ···················· | ···················· |
| T | 1814 | hCoV-19/England/QEUH-B697C9/2020\|EPI_ISL_661423\|20 | ···················· | ···················· | ···················· | ···················· | ···················· | ···················· |
| T | 1815 | hCoV-19/England/QEUH-B67EB7/2020\|EPI_ISL_661424\|20 | ···················· | ···················· | ···················· | ···················· | ···················· | ···················· |
| T | 1816 | hCoV-19/England/QEUH-B666B0/2020\|EPI_ISL_661425\|20 | ···················· | ···················· | ···················· | ···················· | ······T············· | ···················· |
| T | 1817 | hCoV-19/England/QEUH-B6C605/2020\|EPI_ISL_661427\|20 | ···················· | ···················· | ···················· | ···················· | ······T············· | ···················· |
| T | 1818 | hCoV-19/England/QEUH-B67B92/2020\|EPI_ISL_661428\|20 | ···················· | ···················· | ···················· | ···················· | ···················· | ···················· |
| T | 1819 | hCoV-19/England/QEUH-B6C517/2020\|EPI_ISL_661429\|20 | ···················· | ···················· | ···················· | ···················· | ···················· | ···················· |
| T | 1820 | hCoV-19/England/QEUH-B67734/2020\|EPI_ISL_661430\|20 | ···················· | ·A·················· | ···················· | ···················· | ···················· | ···A················ |
| T | 1821 | hCoV-19/England/QEUH-B6778F/2020\|EPI_ISL_661431\|20 | ···················· | ···················· | ···················· | ···················· | ···················· | ···················· |
| T | 1822 | hCoV-19/England/QEUH-B6984E/2020\|EPI_ISL_661432\|20 | ···················· | ···················· | ···················· | ···················· | ···················· | ···················· |
| T | 1823 | hCoV-19/England/QEUH-B6978D/2020\|EPI_ISL_661435\|20 | ···················· | ···················· | ···················· | ···················· | ···················· | ···················· |
| T | 1824 | hCoV-19/England/QEUH-B699F0/2020\|EPI_ISL_661436\|20 | ···················· | ···················· | ···················· | ···················· | ······T············· | ···················· |
| T | 1825 | hCoV-19/England/QEUH-B67B65/2020\|EPI_ISL_661437\|20 | ···················· | ···················· | ···················· | ···················· | ···················· | ···················· |
| T | 1826 | hCoV-19/England/QEUH-B67E99/2020\|EPI_ISL_661438\|20 | ···················· | ···················· | ···················· | ···················· | ···················· | ···················· |
| T | 1827 | hCoV-19/England/QEUH-B665B3/2020\|EPI_ISL_661439\|20 | ···················· | ···················· | ···················· | ···················· | ···················· | ···················· |
| T | 1828 | hCoV-19/England/QEUH-B67637/2020\|EPI_ISL_661440\|20 | ···················· | ·A·················· | ···················· | ···················· | ···················· | ···················· |
| T | 1829 | hCoV-19/England/QEUH-B67AD1/2020\|EPI_ISL_661441\|20 | ···················· | ···················· | ···················· | ···················· | ···················· | ···················· |
| T | 1830 | hCoV-19/England/QEUH-B68F59/2020\|EPI_ISL_661443\|20 | ···················· | ···················· | ···················· | ···················· | ···················· | ···················· |
| T | 1831 | hCoV-19/England/QEUH-B683F3/2020\|EPI_ISL_661444\|20 | ···················· | ···················· | ···················· | ···················· | ···················· | ···················· |
| T | 1832 | hCoV-19/England/QEUH-B66234/2020\|EPI_ISL_661445\|20 | ···················· | ····A··············· | ···················· | ···················· | ···················· | ···················· |
| T | 1833 | hCoV-19/England/QEUH-B67BED/2020\|EPI_ISL_661446\|20 | ···················· | ···················· | ···················· | ···················· | ···················· | ···················· |
| T | 1834 | hCoV-19/England/QEUH-B699E1/2020\|EPI_ISL_661448\|20 | ···················· | ···················· | ···················· | ···················· | ···················· | ···················· |
| T | 1835 | hCoV-19/England/QEUH-B67B47/2020\|EPI_ISL_661449\|20 | ···················· | ···················· | ···················· | ···················· | ····T··············· | ···················· |
| T | 1836 | hCoV-19/England/QEUH-B67743/2020\|EPI_ISL_661454\|20 | ···················· | ···················· | ···················· | ···················· | ···················· | ···················· |
| T | 1837 | hCoV-19/England/QEUH-B676EC/2020\|EPI_ISL_661455\|20 | ···················· | ···················· | ···················· | ···················· | ···················· | ···················· |
| T | 1838 | hCoV-19/England/QEUH-B693E3/2020\|EPI_ISL_661456\|20 | ···················· | ···················· | ···················· | ···················· | ···················· | ···················· |
| T | 1839 | hCoV-19/England/QEUH-B6828D/2020\|EPI_ISL_661457\|20 | ···················· | ···················· | ···················· | ···················· | ···················· | ···················· |
| T | 1840 | hCoV-19/England/QEUH-B697F6/2020\|EPI_ISL_661459\|20 | ···················· | ···················· | ···················· | ···················· | ···················· | ···················· |
| T | 1841 | hCoV-19/England/QEUH-B69310/2020\|EPI_ISL_661460\|20 | ···················· | ·A·················· | ···················· | ···················· | ···················· | ···················· |
| T | 1842 | hCoV-19/England/QEUH-B67664/2020\|EPI_ISL_661462\|20 | ···················· | ···················· | ···················· | ···················· | ···················· | ···················· |
| T | 1843 | hCoV-19/England/QEUH-B66F97/2020\|EPI_ISL_661463\|20 | ···················· | ···················· | ···················· | ···················· | ···················· | ···················· |
| T | 1844 | hCoV-19/England/QEUH-B69750/2020\|EPI_ISL_661464\|20 | ···················· | ···················· | ···················· | ···················· | ···················· | ···················· |
| T | 1845 | hCoV-19/England/QEUH-B69A1B/2020\|EPI_ISL_661466\|20 | ···················· | ···················· | ···················· | ···················· | ···················· | ···················· |
| T | 1846 | hCoV-19/England/QEUH-B67682/2020\|EPI_ISL_661468\|20 | ···················· | ···················· | ···················· | ···················· | ···················· | ···················· |
| T | 1847 | hCoV-19/England/QEUH-B6977E/2020\|EPI_ISL_661469\|20 | ···················· | ···················· | ···················· | ···················· | ···················· | ···················· |
| T | 1848 | hCoV-19/England/QEUH-B697BA/2020\|EPI_ISL_661470\|20 | ···················· | ···················· | ···················· | ···················· | ···················· | ···················· |
| T | 1849 | hCoV-19/England/QEUH-B689A6/2020\|EPI_ISL_661471\|20 | ···················· | ·A·················· | ···················· | ···················· | ···················· | ···A················ |
| T | 1850 | hCoV-19/England/QEUH-B6837B/2020\|EPI_ISL_661472\|20 | ···················· | ···················· | ···················· | ···················· | ···················· | ···················· |
| T | 1851 | hCoV-19/England/QEUH-B67646/2020\|EPI_ISL_661475\|20 | ···················· | ···················· | ···················· | ···················· | ···················· | ···················· |
| T | 1852 | hCoV-19/England/QEUH-B6995A/2020\|EPI_ISL_661476\|20 | ···················· | ···················· | ···················· | ···················· | ······T············· | ···················· |
| T | 1853 | hCoV-19/England/QEUH-B6932F/2020\|EPI_ISL_661477\|20 | ···················· | ···················· | ···················· | ···················· | ···················· | ···················· |
| T | 1854 | hCoV-19/England/QEUH-B66489/2020\|EPI_ISL_661478\|20 | ···················· | ···················· | ···················· | ···················· | ···················· | ···················· |
| T | 1855 | hCoV-19/England/QEUH-B69AA2/2020\|EPI_ISL_661480\|20 | ···················· | ···················· | ···················· | ···················· | ···················· | ···················· |
| T | 1856 | hCoV-19/England/QEUH-B663AA/2020\|EPI_ISL_661481\|20 | ···················· | ···················· | ···················· | ···················· | ···················· | ···················· |
| T | 1857 | hCoV-19/England/QEUH-B67C08/2020\|EPI_ISL_661482\|20 | ···················· | ···················· | ···················· | ···················· | ···················· | ···················· |
| T | 1858 | hCoV-19/England/QEUH-B6933E/2020\|EPI_ISL_661483\|20 | ···················· | ···················· | ···················· | ···················· | ···················· | ···················· |
| T | 1859 | hCoV-19/England/QEUH-B6992D/2020\|EPI_ISL_661484\|20 | ···················· | ···················· | ···················· | ···················· | ···················· | ···················· |
| T | 1860 | hCoV-19/England/QEUH-B6701B/2020\|EPI_ISL_661486\|20 | ···················· | ···················· | ···················· | ···················· | ···················· | ···················· |
| T | 1861 | hCoV-19/England/QEUH-B67AFF/2020\|EPI_ISL_661487\|20 | ···················· | ···················· | ···················· | ···················· | ···················· | ···················· |
| T | 1862 | hCoV-19/England/QEUH-B69B09/2020\|EPI_ISL_661489\|20 | ···················· | ···················· | ···················· | ···················· | ···················· | ···················· |
| T | 1863 | hCoV-19/England/QEUH-B66313/2020\|EPI_ISL_661490\|20 | ···················· | ···················· | ···················· | ···················· | ···················· | ···················· |
| T | 1864 | hCoV-19/England/QEUH-B6838A/2020\|EPI_ISL_661492\|20 | ···················· | ···················· | ···················· | ···················· | ···················· | ···················· |
| T | 1865 | hCoV-19/England/QEUH-B67B83/2020\|EPI_ISL_661493\|20 | ···················· | ···················· | ···················· | ···················· | ···················· | ···················· |
| T | 1866 | hCoV-19/England/QEUH-B699A5/2020\|EPI_ISL_661496\|20 | ···················· | ···················· | ···················· | ···················· | ···················· | ···················· |
| T | 1867 | hCoV-19/England/QEUH-B682AB/2020\|EPI_ISL_661497\|20 | ···················· | ···················· | ···················· | ···················· | ···················· | ···················· |
| T | 1868 | hCoV-19/England/QEUH-B676DD/2020\|EPI_ISL_661498\|20 | ···················· | ···················· | ···················· | ···················· | ···················· | ···················· |

T 1869 hCoV-19/England/QEUH-AF05DD/2020|EPI_ISL_662701|20 ···················· ·A·················· ···················· ···················· ···················· ··A·················
T 1870 hCoV-19/England/QEUH-AF2948/2020|EPI_ISL_662702|20 ···················· ···················· ···················· ···················· ···················· ····················
T 1871 hCoV-19/England/QEUH-AF018E/2020|EPI_ISL_662703|20 ···················· ···················· ···················· ···················· ···················· ····················
T 1872 hCoV-19/England/QEUH-AEFB93/2020|EPI_ISL_662704|20 ···················· ···················· ···················· ···················· ···················· ····················
T 1873 hCoV-19/England/QEUH-AF0BF9/2020|EPI_ISL_662705|20 ···················· ···················· ···················· ···················· ···················· ····················
T 1874 hCoV-19/England/QEUH-AEFC45/2020|EPI_ISL_662706|20 ···················· ···················· ···················· ···················· ···················· ····················
T 1875 hCoV-19/England/QEUH-AEFB57/2020|EPI_ISL_662707|20 ···················· ···················· ···················· ···················· ···················· ····················
T 1876 hCoV-19/England/QEUH-AF0C23/2020|EPI_ISL_662710|20 ···················· ···················· ···················· ···················· ···················· ····················
T 1877 hCoV-19/England/QEUH-AF25F9/2020|EPI_ISL_662711|20 ···················· ···················· ···················· ···················· ···················· ····················
T 1878 hCoV-19/England/QEUH-AF1017/2020|EPI_ISL_662713|20 ···················· ···················· ···················· ···················· ···················· ····················
T 1879 hCoV-19/England/QEUH-AEFD51/2020|EPI_ISL_662715|20 ···················· ···················· ···················· ···················· ···················· ····················
T 1880 hCoV-19/England/QEUH-AF0106/2020|EPI_ISL_662716|20 ···················· ···················· ···················· ···················· ·····T·············· ····················
T 1881 hCoV-19/England/QEUH-AEFDCA/2020|EPI_ISL_662720|20 ···················· ···················· ···················· ···················· ···················· ····················
T 1882 hCoV-19/England/QEUH-AF00EB/2020|EPI_ISL_662724|20 ···················· ···················· ···················· ···················· ···················· ····················
T 1883 hCoV-19/England/QEUH-AEFD24/2020|EPI_ISL_662726|20 ···················· ···················· ···················· ···················· ···················· ····················
T 1884 hCoV-19/England/QEUH-AF291B/2020|EPI_ISL_662727|20 ···················· ···················· ···················· ···················· ···················· ····················
T 1885 hCoV-19/England/QEUH-AEFD06/2020|EPI_ISL_662729|20 ···················· ·A·················· ···················· ···················· ···················· ····················
T 1886 hCoV-19/England/QEUH-AF473D/2020|EPI_ISL_662730|20 ···················· ···················· ···················· ···················· ···················· ····················
T 1887 hCoV-19/England/QEUH-AF2562/2020|EPI_ISL_662731|20 ···················· ···················· ···················· ···················· ···················· ····················
T 1888 hCoV-19/England/QEUH-AF075F/2020|EPI_ISL_662732|20 ···················· ·A·················· ···················· ···················· ···················· ··A·················
T 1889 hCoV-19/England/QEUH-AF2632/2020|EPI_ISL_662734|20 ···················· ···················· ···················· ···················· ···················· ····················
T 1890 hCoV-19/England/QEUH-AEF98A/2020|EPI_ISL_662736|20 ···················· ·A·················· ···················· ···················· ···················· ··A·················
T 1891 hCoV-19/England/QEUH-AF290C/2020|EPI_ISL_662737|20 ···················· ···················· ···················· ···················· ···················· ····················
T 1892 hCoV-19/England/QEUH-AF27D5/2020|EPI_ISL_662739|20 ···················· ···················· ···················· ···················· ···················· ····················
T 1893 hCoV-19/England/QEUH-AF05EC/2020|EPI_ISL_662746|20 ···················· ···················· ···················· ···················· ···················· ····················
T 1894 hCoV-19/England/QEUH-AEFAC3/2020|EPI_ISL_662747|20 ···················· ···················· ···················· ···················· ···················· ····················
T 1895 hCoV-19/England/QEUH-AEFC90/2020|EPI_ISL_662748|20 ···················· ···················· ···················· ···················· ···················· ····················
T 1896 hCoV-19/England/QEUH-AF00BE/2020|EPI_ISL_662749|20 ···················· ···················· ···················· ···················· ···················· ····················
T 1897 hCoV-19/England/QEUH-AF2939/2020|EPI_ISL_662750|20 ···················· ···················· ···················· ···················· ···················· ····················
T 1898 hCoV-19/England/QEUH-AEFD33/2020|EPI_ISL_662752|20 ···················· ···················· ···················· ···················· ···················· ····················
T 1899 hCoV-19/England/QEUH-AF28F0/2020|EPI_ISL_662753|20 ···················· ···················· ···················· ···················· ···················· ····················
T 1900 hCoV-19/England/QEUH-AF0C05/2020|EPI_ISL_662754|20 ···················· ···················· ···················· ···················· ···················· ····················
T 1901 hCoV-19/England/QEUH-AEF9C6/2020|EPI_ISL_662755|20 ···················· ···················· ···················· ···················· ···················· ····················
T 1902 hCoV-19/England/QEUH-AEFE12/2020|EPI_ISL_662756|20 ···················· ···················· ···················· ···················· ···················· ····················
T 1903 hCoV-19/England/QEUH-AF0FDF/2020|EPI_ISL_662757|20 ···················· ···················· ···················· ···················· ···················· ····················
T 1904 hCoV-19/England/QEUH-AEFCCD/2020|EPI_ISL_662758|20 ···················· ···················· ···················· ···················· ···················· ····················
T 1905 hCoV-19/England/QEUH-AF0731/2020|EPI_ISL_662760|20 ···················· ···················· ···················· ···················· ···················· ····················
T 1906 hCoV-19/England/QEUH-AF0713/2020|EPI_ISL_662761|20 ···················· ···················· ···················· ···················· ···················· ····················
T 1907 hCoV-19/England/QEUH-AF0959/2020|EPI_ISL_662764|20 ···················· ···················· ···················· ···················· ···················· ····················
T 1908 hCoV-19/England/QEUH-AEFF1F/2020|EPI_ISL_662765|20 ···················· ···················· ···················· ···················· ···················· ····················
T 1909 hCoV-19/England/QEUH-AEFFD3/2020|EPI_ISL_662766|20 ···················· ···················· ···················· ···················· ···················· ····················
T 1910 hCoV-19/England/QEUH-AF068F/2020|EPI_ISL_662767|20 ···················· ···················· ···················· ···················· ···················· ····················
T 1911 hCoV-19/England/QEUH-AEFDF7/2020|EPI_ISL_662768|20 ···················· ···················· ···················· ···················· ···················· ····················
T 1912 hCoV-19/England/QEUH-AF23A4/2020|EPI_ISL_662769|20 ···················· ···················· ···················· ···················· ···················· ····················
T 1913 hCoV-19/England/QEUH-AF109F/2020|EPI_ISL_662771|20 ···················· ···················· ···················· ···················· ···················· ····················
T 1914 hCoV-19/England/QEUH-AF285A/2020|EPI_ISL_662775|20 ···················· ···················· ···················· ···················· ···················· ····················
T 1915 hCoV-19/England/QEUH-AEFCDC/2020|EPI_ISL_662776|20 ···················· ···················· ···················· ···················· ···················· ····················
T 1916 hCoV-19/England/QEUH-AEFB1B/2020|EPI_ISL_662777|20 ···················· ···················· ···················· ···················· ···················· ····················
T 1917 hCoV-19/England/QEUH-AEFB48/2020|EPI_ISL_662778|20 ···················· ···················· ···················· ···················· ···················· ····················
T 1918 hCoV-19/England/QEUH-AF0537/2020|EPI_ISL_662779|20 ···················· ···················· ···················· ···················· ···················· ····················
T 1919 hCoV-19/England/QEUH-AEFD9D/2020|EPI_ISL_662785|20 ···················· ···················· ···················· ···················· ···················· ····················
T 1920 hCoV-19/England/QEUH-AF0CD8/2020|EPI_ISL_662787|20 ···················· ···················· ···················· ···················· ···················· ····················
T 1921 hCoV-19/England/QEUH-AF0BDB/2020|EPI_ISL_662789|20 ···················· ···················· ···················· ···················· ···················· ····················
T 1922 hCoV-19/England/QEUH-AEF9B7/2020|EPI_ISL_662791|20 ···················· ···················· ···················· ···················· ···················· ····················
T 1923 hCoV-19/England/QEUH-AF00DC/2020|EPI_ISL_662792|20 ···················· ···················· ···················· ···················· ···················· ····················
T 1924 hCoV-19/England/QEUH-AF0CE7/2020|EPI_ISL_662794|20 ···················· ···················· ···················· ···················· ···················· ····················
T 1925 hCoV-19/England/QEUH-AF0FB1/2020|EPI_ISL_662795|20 ···················· ···················· ···················· ···················· ···················· ····················
T 1926 hCoV-19/England/QEUH-AEFD7F/2020|EPI_ISL_662796|20 ···················· ···················· ···················· ···················· ···················· ····················
T 1927 hCoV-19/England/QEUH-AF06CB/2020|EPI_ISL_662797|20 ···················· ···················· ···················· ···················· ···················· ····················
T 1928 hCoV-19/England/CAMB-1BB416/2020|EPI_ISL_665007|20 ···················· ···················· ···················· ···················· ···················· ····················
T 1929 hCoV-19/England/CAMB-1BB443/2020|EPI_ISL_665008|20 ···················· ···················· ···················· ···················· ···················· ····················
T 1930 hCoV-19/England/CAMB-1BB452/2020|EPI_ISL_665009|20 ···················· ···················· ···················· ···················· ···T················ ····················
T 1931 hCoV-19/England/CAMB-1BB49E/2020|EPI_ISL_665010|20 ···················· ···················· ···················· ···················· ···················· ····················
T 1932 hCoV-19/England/CAMB-1BB59B/2020|EPI_ISL_665011|20 ···················· ···················· ···················· ···················· ···················· ····················
T 1934 hCoV-19/England/CAMB-1BB5E6/2020|EPI_ISL_665013|20 ···················· ···················· ···················· ···················· ···················· ····················
T 1935 hCoV-19/England/CAMB-1BB610/2020|EPI_ISL_665014|20 ···················· ···················· ···················· ···················· ···················· ····················
T 1936 hCoV-19/England/CAMB-1BB62F/2020|EPI_ISL_665015|20 ···················· ···················· ···················· ···················· ···················· ····················
T 1938 hCoV-19/England/CAMB-1BB689/2020|EPI_ISL_665017|20 ···················· ···················· ···················· ···················· ···················· ····················
T 1939 hCoV-19/England/CAMC-B2301B/2020|EPI_ISL_665025|20 ···················· ···················· ···················· ···················· ···················· ····················
T 1940 hCoV-19/England/CAMC-B23048/2020|EPI_ISL_665027|20 ···················· ···················· ···················· ···················· ···················· ····················
T 1941 hCoV-19/England/CAMC-B231DC/2020|EPI_ISL_665028|20 ···················· ·A·················· ···················· ···················· ···················· ··A·················
T 1942 hCoV-19/Wales/PHWC-48BC41/2020|EPI_ISL_666338|2020 ···················· ···················· ···················· ···················· ···················· ····················
T 1943 hCoV-19/Wales/PHWC-48BC50/2020|EPI_ISL_666339|2020 ···················· ···················· ···················· ···················· ···················· ····················
T 1944 hCoV-19/Wales/PHWC-48BC6F/2020|EPI_ISL_666340|2020 ···················· ···················· ···················· ···················· ···················· ····················
T 1945 hCoV-19/Wales/PHWC-48BC8D/2020|EPI_ISL_666341|2020 ···················· ···················· ···················· ···················· ···················· ····················
T 1946 hCoV-19/Wales/PHWC-48BC9C/2020|EPI_ISL_666342|2020 ···················· ···················· ···················· ···················· ···················· ····················
T 1947 hCoV-19/Wales/PHWC-48BCAB/2020|EPI_ISL_666343|2020 ···················· ···················· ···················· ···················· ···················· ····················
T 1948 hCoV-19/Wales/PHWC-48BCBA/2020|EPI_ISL_666344|2020 ···················· ···················· ···················· ···················· ···················· ····················
T 1949 hCoV-19/Wales/PHWC-48BCC9/2020|EPI_ISL_666345|2020 ···················· ···················· ···················· ···················· ···················· ····················
T 1950 hCoV-19/Wales/PHWC-48BCF6/2020|EPI_ISL_666346|2020 ···················· ···················· ···················· ···················· ···················· ····················

| | | | | | | | |
|---|---|---|---|---|---|---|---|
| T | 1951 hCoV-19/Wales/PHWC-48BD02/2020\|EPI_ISL_666347\|2020 | ···················· | ···················· | ···················· | ···················· | ···················· | ···················· |
| T | 1952 hCoV-19/Wales/PHWC-48BD11/2020\|EPI_ISL_666348\|2020 | ···················· | ···················· | ···················· | ···················· | ···················· | ···················· |
| T | 1953 hCoV-19/Wales/PHWC-48BDA8/2020\|EPI_ISL_666356\|2020 | ···················· | ···················· | ···················· | ···················· | ···················· | ···················· |
| T | 1954 hCoV-19/Wales/PHWC-48BDB7/2020\|EPI_ISL_666357\|2020 | ···················· | ···················· | ···················· | ···················· | ···················· | ···················· |
| T | 1955 hCoV-19/Wales/PHWC-48BDD5/2020\|EPI_ISL_666359\|2020 | ···················· | ···················· | ···················· | ···················· | ···················· | ···················· |
| T | 1956 hCoV-19/Japan/IC-0240/2020\|EPI_ISL_667669\|2020-10 | ···················· | ···················· | ···················· | ···················· | ···················· | ···················· |
| T | 1957 hCoV-19/Japan/IC-0241/2020\|EPI_ISL_667670\|2020-10 | ···················· | ···················· | ···················· | ···················· | ···················· | ···················· |
| T | 1958 hCoV-19/Japan/IC-0242/2020\|EPI_ISL_667671\|2020-10 | ···················· | ···················· | ···················· | ···················· | ·····T·············· | ···················· |
| T | 1959 hCoV-19/Japan/IC-0243/2020\|EPI_ISL_667672\|2020-10 | ···················· | ·A·················· | ···················· | ···················· | ···················· | ···A················ |
| T | 1960 hCoV-19/Japan/IC-0244/2020\|EPI_ISL_667673\|2020-10 | ···················· | ···················· | ···················· | ···················· | ···················· | ···················· |
| T | 1961 hCoV-19/Japan/IC-0245/2020\|EPI_ISL_667674\|2020-10 | ···················· | ·A·················· | ···················· | ···················· | ···················· | ···A················ |
| T | 1962 hCoV-19/Japan/IC-0246/2020\|EPI_ISL_667675\|2020-10 | ···················· | ·A·················· | ···················· | ···················· | ···················· | ···A················ |
| T | 1963 hCoV-19/Japan/IC-0247/2020\|EPI_ISL_667676\|2020-10 | ···················· | ···················· | ···················· | ···················· | ···················· | ···················· |
| T | 1964 hCoV-19/Japan/IC-0248/2020\|EPI_ISL_667677\|2020-10 | ···················· | ···················· | ···················· | ···················· | ···················· | ···················· |
| T | 1965 hCoV-19/Japan/IC-0249/2020\|EPI_ISL_667678\|2020-10 | ···················· | ···················· | ···················· | ···················· | ···················· | ···················· |
| T | 1966 hCoV-19/Japan/IC-0250/2020\|EPI_ISL_667679\|2020-10 | ···················· | ···················· | ···················· | ···················· | ···················· | ···················· |
| T | 1967 hCoV-19/Japan/IC-0258/2020\|EPI_ISL_667687\|2020-10 | ···················· | ···················· | ···················· | ···················· | ···················· | ···················· |
| T | 1968 hCoV-19/Japan/IC-0259/2020\|EPI_ISL_667688\|2020-10 | ···················· | ···················· | ···················· | ···················· | ···················· | ···················· |
| T | 1969 hCoV-19/Denmark/DCGC-9306/2020\|EPI_ISL_668900\|2020 | ···················· | ···················· | ···················· | ···················· | ···················· | ···················· |
| T | 1970 hCoV-19/Denmark/DCGC-9309/2020\|EPI_ISL_668901\|2020 | ···················· | ···················· | ···················· | ···················· | ···················· | ···················· |
| T | 1971 hCoV-19/Denmark/DCGC-9312/2020\|EPI_ISL_668902\|2020 | ···················· | ···················· | ···················· | ···················· | ···················· | ···················· |
| T | 1972 hCoV-19/Denmark/DCGC-9314/2020\|EPI_ISL_668903\|2020 | ···················· | ···················· | ···················· | ···················· | ···················· | ···················· |
| T | 1973 hCoV-19/Denmark/DCGC-9315/2020\|EPI_ISL_668904\|2020 | ···················· | ···················· | ···················· | ···················· | ···················· | ···················· |
| T | 1974 hCoV-19/Denmark/DCGC-9319/2020\|EPI_ISL_668905\|2020 | ···················· | ···················· | ···················· | ···················· | ···················· | ···················· |
| T | 1975 hCoV-19/Denmark/DCGC-9320/2020\|EPI_ISL_668906\|2020 | ···················· | ···················· | ···················· | ···················· | ···················· | ···················· |
| T | 1976 hCoV-19/Denmark/DCGC-9322/2020\|EPI_ISL_668907\|2020 | ···················· | ···················· | ···················· | ···················· | ···················· | ···················· |
| T | 1977 hCoV-19/Denmark/DCGC-9326/2020\|EPI_ISL_668908\|2020 | ···················· | ···················· | ···················· | ···················· | ···················· | ···················· |
| T | 1978 hCoV-19/Denmark/DCGC-9328/2020\|EPI_ISL_668909\|2020 | ···················· | ···················· | ···················· | ···················· | ···················· | ···················· |
| T | 1979 hCoV-19/Denmark/DCGC-9329/2020\|EPI_ISL_668910\|2020 | ···················· | ···················· | ···················· | ···················· | ···················· | ···················· |
| T | 1980 hCoV-19/Denmark/DCGC-9330/2020\|EPI_ISL_668911\|2020 | ···················· | ···················· | ···················· | ···················· | ···················· | ···················· |
| T | 1982 hCoV-19/USA/CA-CZB-10121/2020\|EPI_ISL_672112\|2020- | ···················· | ···················· | ···················· | ···················· | ···················· | ···················· |
| T | 1983 hCoV-19/USA/CA-CZB-13412/2020\|EPI_ISL_672113\|2020- | ···················· | ···················· | ···················· | ···················· | ···················· | ···················· |
| T | 1984 hCoV-19/USA/CA-CZB-9518/2020\|EPI_ISL_672114\|2020-0 | ···················· | ···················· | ···················· | ···················· | ···················· | ···················· |
| T | 1985 hCoV-19/USA/CA-CZB-9519/2020\|EPI_ISL_672115\|2020-0 | ···················· | ···················· | ···················· | ···················· | ···················· | ···················· |
| T | 1986 hCoV-19/USA/CA-CZB-9520/2020\|EPI_ISL_672116\|2020-0 | ···················· | ···················· | ···················· | ···················· | ···················· | ···················· |
| T | 1987 hCoV-19/USA/CA-CZB-9522/2020\|EPI_ISL_672117\|2020-0 | ···················· | ···················· | ···················· | ···················· | ···················· | ···················· |
| T | 1988 hCoV-19/USA/CA-CZB-9523/2020\|EPI_ISL_672118\|2020-0 | ···················· | ···················· | ···················· | ···················· | ···················· | ···················· |
| T | 1990 hCoV-19/USA/CA-CZB-9527/2020\|EPI_ISL_672120\|2020-0 | ···················· | ···················· | ···················· | ···················· | ···················· | ···················· |
| T | 1991 hCoV-19/USA/CA-CZB-9529/2020\|EPI_ISL_672121\|2020-0 | ···················· | ···················· | ···················· | ···················· | ···················· | ···················· |
| T | 1992 hCoV-19/USA/CA-CZB-9533/2020\|EPI_ISL_672122\|2020-0 | ···················· | ···················· | ···················· | ···················· | ···················· | ···················· |
| T | 1993 hCoV-19/USA/CA-CZB-9539/2020\|EPI_ISL_672123\|2020-0 | ···················· | ···················· | ···················· | ···················· | ···················· | ···················· |
| T | 1994 hCoV-19/USA/CA-CZB-9540/2020\|EPI_ISL_672124\|2020-0 | ···················· | ···················· | ···················· | ···················· | ···················· | ···················· |
| T | 1997 hCoV-19/USA/CA-CZB-9547/2020\|EPI_ISL_672127\|2020-0 | ···················· | ···················· | ···················· | ···················· | ···················· | ···················· |
| T | 1998 hCoV-19/USA/CA-CZB-9548/2020\|EPI_ISL_672128\|2020-0 | ···················· | ···················· | ···················· | ···················· | ···················· | ···················· |
| T | 1999 hCoV-19/USA/CA-CZB-9549/2020\|EPI_ISL_672129\|2020-0 | ···················· | ···················· | ···················· | ···················· | ···················· | ···················· |
| T | 2000 hCoV-19/USA/CA-CZB-9552/2020\|EPI_ISL_672130\|2020-0 | ···················· | ···················· | ···················· | ···················· | ···················· | ···················· |
| T | 2001 hCoV-19/USA/CA-CZB-9556/2020\|EPI_ISL_672131\|2020-0 | ···················· | ···················· | ···················· | ···················· | ···················· | ···················· |
| T | 2002 hCoV-19/USA/CA-CZB-9560/2020\|EPI_ISL_672132\|2020-0 | ···················· | ···················· | ···················· | ···················· | ···················· | ···················· |
| T | 2003 hCoV-19/USA/CA-CZB-9563/2020\|EPI_ISL_672133\|2020-0 | ···················· | ···················· | ···················· | ···················· | ···················· | ···················· |
| T | 2004 hCoV-19/USA/CA-CZB-9571/2020\|EPI_ISL_672134\|2020-0 | ···················· | ···················· | ···················· | ···················· | ···················· | ···················· |
| T | 2005 hCoV-19/USA/CA-CZB-9574/2020\|EPI_ISL_672135\|2020-0 | ···················· | ···················· | ···················· | ···················· | ···················· | ···················· |
| T | 2006 hCoV-19/USA/CA-CZB-9585/2020\|EPI_ISL_672136\|2020-0 | ···················· | ···················· | ···················· | ···················· | ···················· | ···················· |
| T | 2007 hCoV-19/USA/CA-CZB-9589/2020\|EPI_ISL_672137\|2020-0 | ···················· | ···················· | ···················· | ···················· | ···················· | ···················· |
| T | 2008 hCoV-19/USA/CA-CZB-9590/2020\|EPI_ISL_672138\|2020-0 | ···················· | ···················· | ···················· | ···················· | ···················· | ···················· |
| T | 2009 hCoV-19/USA/CA-CZB-9597/2020\|EPI_ISL_672139\|2020-0 | ···················· | ···················· | ···················· | ···················· | ···················· | ···················· |
| T | 2010 hCoV-19/USA/CA-CZB-9610/2020\|EPI_ISL_672140\|2020-0 | ···················· | ···················· | ···················· | ···················· | ···················· | ···················· |
| T | 2011 hCoV-19/USA/CA-CZB-9614/2020\|EPI_ISL_672141\|2020-0 | ···················· | ···················· | ···················· | ···················· | ···················· | ···················· |
| T | 2012 hCoV-19/USA/CA-CZB-9625/2020\|EPI_ISL_672142\|2020-0 | ···················· | ···················· | ···················· | ···················· | ···················· | ···················· |
| T | 2013 hCoV-19/USA/CA-CZB-9631/2020\|EPI_ISL_672143\|2020-0 | ···················· | ···················· | ···················· | ···················· | ···················· | ···················· |
| T | 2014 hCoV-19/USA/CA-CZB-9638/2020\|EPI_ISL_672144\|2020-0 | ···················· | ···················· | ···················· | ···················· | ···················· | ···················· |
| T | 2015 hCoV-19/USA/CA-CZB-9640/2020\|EPI_ISL_672145\|2020-0 | ···················· | ···················· | ···················· | ···················· | ···················· | ···················· |
| T | 2016 hCoV-19/USA/CA-CZB-9645/2020\|EPI_ISL_672146\|2020-0 | ···················· | ···················· | ···················· | ···················· | ···················· | ···················· |
| T | 2017 hCoV-19/USA/CA-CZB-9647/2020\|EPI_ISL_672147\|2020-0 | ···················· | ···················· | ···················· | ···················· | ···················· | ···················· |
| T | 2018 hCoV-19/USA/CA-CZB-9648/2020\|EPI_ISL_672148\|2020-0 | ···················· | ···················· | ···················· | ···················· | ···················· | ···················· |
| T | 2019 hCoV-19/USA/CA-CZB-9650/2020\|EPI_ISL_672149\|2020-0 | ···················· | ···················· | ···················· | ···················· | ···················· | ···················· |
| T | 2020 hCoV-19/USA/CA-CZB-9653/2020\|EPI_ISL_672150\|2020-0 | ···················· | ···················· | ···················· | ···················· | ···················· | ···················· |
| T | 2021 hCoV-19/USA/CA-CZB-9656/2020\|EPI_ISL_672151\|2020-0 | ···················· | ···················· | ···················· | ···················· | ···················· | ···················· |
| T | 2022 hCoV-19/USA/CA-CZB-9657/2020\|EPI_ISL_672152\|2020-0 | ···················· | ···················· | ···················· | ···················· | ···················· | ···················· |
| T | 2023 hCoV-19/USA/CA-CZB-9675/2020\|EPI_ISL_672153\|2020-0 | ···················· | ···················· | ···················· | ···················· | ···················· | ···················· |
| T | 2024 hCoV-19/USA/CA-CZB-9682/2020\|EPI_ISL_672154\|2020-0 | ···················· | ···················· | ···················· | ···················· | ···················· | ···················· |
| T | 2025 hCoV-19/USA/CA-CZB-9705/2020\|EPI_ISL_672155\|2020-0 | ···················· | ···················· | ···················· | ···················· | ···················· | ···················· |
| T | 2028 hCoV-19/USA/CA-CZB-9716/2020\|EPI_ISL_672158\|2020-0 | ···················· | ···················· | ···················· | ···················· | ···················· | ···················· |
| T | 2029 hCoV-19/USA/CA-CZB-9717/2020\|EPI_ISL_672159\|2020-0 | ···················· | ···················· | ···················· | ···················· | ···················· | ···················· |
| T | 2030 hCoV-19/USA/CA-CZB-9724/2020\|EPI_ISL_672160\|2020-0 | ···················· | ···················· | ···················· | ···················· | ···················· | ···················· |
| T | 2031 hCoV-19/USA/CA-CZB-9729/2020\|EPI_ISL_672161\|2020-0 | ···················· | ···················· | ···················· | ···················· | ···················· | ···················· |
| T | 2032 hCoV-19/USA/CA-CZB-9731/2020\|EPI_ISL_672162\|2020-0 | ···················· | ···················· | ···················· | ···················· | ···················· | ···················· |
| T | 2033 hCoV-19/USA/CA-CZB-9750/2020\|EPI_ISL_672163\|2020-0 | ···················· | ···················· | ···················· | ···················· | ···················· | ···················· |
| T | 2034 hCoV-19/USA/CA-CZB-9751/2020\|EPI_ISL_672164\|2020-0 | ···················· | ···················· | ···················· | ···················· | ···················· | ···················· |
| T | 2035 hCoV-19/USA/CA-CZB-9753/2020\|EPI_ISL_672165\|2020-0 | ···················· | ···················· | ···················· | ···················· | ···················· | ···················· |
| T | 2036 hCoV-19/USA/CA-CZB-9754/2020\|EPI_ISL_672166\|2020-0 | ···················· | ···················· | ···················· | ···················· | ···················· | ···················· |

| | | | | | | | |
|---|---|---|---|---|---|---|---|
| T | 2037 hCoV-19/USA/CA-CZB-9756/2020\|EPI_ISL_672167\|2020-0 | ···················· | ···················· | ···················· | ···················· | ···················· | ···················· |
| T | 2038 hCoV-19/USA/CA-CZB-9759/2020\|EPI_ISL_672168\|2020-0 | ···················· | ···················· | ···················· | ···················· | ···················· | ···················· |
| T | 2039 hCoV-19/USA/CA-CZB-9760/2020\|EPI_ISL_672169\|2020-0 | ···················· | ···················· | ···················· | ···················· | ···················· | ···················· |
| T | 2040 hCoV-19/USA/CA-CZB-9762/2020\|EPI_ISL_672170\|2020-0 | ···················· | ···················· | ···················· | ···················· | ···················· | ···················· |
| T | 2041 hCoV-19/USA/CA-CZB-9768/2020\|EPI_ISL_672171\|2020-0 | ···················· | ···················· | ···················· | ···················· | ···················· | ···················· |
| T | 2042 hCoV-19/USA/CA-CZB-9773/2020\|EPI_ISL_672172\|2020-0 | ···················· | ····A··············· | ···················· | ···················· | ···················· | ···················· |
| T | 2043 hCoV-19/USA/CA-CZB-9774/2020\|EPI_ISL_672173\|2020-0 | ···················· | ···················· | ···················· | ···················· | ···················· | ···················· |
| T | 2044 hCoV-19/USA/CA-CZB-9775/2020\|EPI_ISL_672174\|2020-0 | ···················· | ·A·················· | ···················· | ···················· | ···················· | ···················· |
| T | 2046 hCoV-19/USA/CA-CZB-9784/2020\|EPI_ISL_672176\|2020-0 | ···················· | ···················· | ···················· | ···················· | ···················· | ···················· |
| T | 2047 hCoV-19/USA/CA-CZB-9789/2020\|EPI_ISL_672177\|2020-0 | ···················· | ···················· | ···················· | ···················· | ···················· | ···················· |
| T | 2048 hCoV-19/USA/CA-CZB-9791/2020\|EPI_ISL_672178\|2020-0 | ···················· | ···················· | ···················· | ···················· | ···················· | ···················· |
| T | 2049 hCoV-19/England/QEUH-B68311/2020\|EPI_ISL_661450\|20 | ···················· | ·A·················· | ···················· | ···················· | ···················· | ···················· |
| T | 2050 hCoV-19/England/QEUH-B67B1A/2020\|EPI_ISL_661451\|20 | ···················· | ···················· | ···················· | ···················· | ···················· | ···················· |
| T | 2051 hCoV-19/England/QEUH-B67A77/2020\|EPI_ISL_661452\|20 | ···················· | ···················· | ···················· | ···················· | ···················· | ···················· |
| T | 2052 hCoV-19/England/QEUH-AF47B5/2020\|EPI_ISL_662780\|20 | ···················· | ···················· | ···················· | ···················· | ···················· | ···················· |
| T | 2053 hCoV-19/England/QEUH-AF24FC/2020\|EPI_ISL_662782\|20 | ···················· | ···················· | ···················· | ···················· | ···················· | ···················· |
| T | 2054 hCoV-19/England/QEUH-AF0018/2020\|EPI_ISL_662783\|20 | ···················· | ···················· | ···················· | ···················· | ···················· | ···················· |
| T | 2055 hCoV-19/England/QEUH-AF2957/2020\|EPI_ISL_662784\|20 | ···················· | ···················· | ···················· | ···················· | ···················· | ···················· |
| T | 2056 hCoV-19/Wales/PHWC-48B18E/2020\|EPI_ISL_666200\|2020 | ···················· | ···················· | ···················· | ···················· | ···················· | ···················· |
| T | 2058 hCoV-19/Wales/PHWC-48B1AC/2020\|EPI_ISL_666202\|2020 | ···················· | ···················· | ···················· | ···················· | ···················· | ···················· |
| T | 2059 hCoV-19/Wales/PHWC-48B1BB/2020\|EPI_ISL_666203\|2020 | ···················· | ···················· | ···················· | ···················· | ···················· | ···················· |
| T | 2060 hCoV-19/Wales/PHWC-48B1CA/2020\|EPI_ISL_666204\|2020 | ···················· | ···················· | ···················· | ···················· | ···················· | ···················· |
| T | 2061 hCoV-19/Wales/PHWC-48B1D9/2020\|EPI_ISL_666205\|2020 | ···················· | ···················· | ···················· | ···················· | ···················· | ···················· |
| T | 2062 hCoV-19/Wales/PHWC-48B1E8/2020\|EPI_ISL_666206\|2020 | ···················· | ···················· | ···················· | ···················· | ···················· | ···················· |
| T | 2064 hCoV-19/Wales/PHWC-48B212/2020\|EPI_ISL_666208\|2020 | ···················· | ···················· | ···················· | ···················· | ···················· | ···················· |
| T | 2065 hCoV-19/Wales/PHWC-48B221/2020\|EPI_ISL_666209\|2020 | ···················· | ···················· | ···················· | ···················· | ···················· | ···················· |
| T | 2066 hCoV-19/Wales/PHWC-48B230/2020\|EPI_ISL_666210\|2020 | ···················· | ···················· | ···················· | ···················· | ···················· | ···················· |
| T | 2067 hCoV-19/Wales/PHWC-48B24F/2020\|EPI_ISL_666211\|2020 | ···················· | ···················· | ···················· | ···················· | ···················· | ···················· |
| T | 2068 hCoV-19/Wales/PHWC-48B25E/2020\|EPI_ISL_666212\|2020 | ···················· | ···················· | ···················· | ···················· | ···················· | ···················· |
| T | 2069 hCoV-19/Wales/PHWC-48B26D/2020\|EPI_ISL_666213\|2020 | ···················· | ···················· | ···················· | ···················· | ···················· | ···················· |
| T | 2070 hCoV-19/Wales/PHWC-48B28B/2020\|EPI_ISL_666214\|2020 | ···················· | ···················· | ···················· | ···················· | ···················· | ···················· |
| T | 2071 hCoV-19/Wales/PHWC-48B29A/2020\|EPI_ISL_666215\|2020 | ···················· | ···················· | ···················· | ···················· | ···················· | ···················· |
| T | 2072 hCoV-19/Wales/PHWC-48B2A9/2020\|EPI_ISL_666216\|2020 | ···················· | ···················· | ···················· | ···················· | ···················· | ···················· |
| T | 2073 hCoV-19/Wales/PHWC-48B2B8/2020\|EPI_ISL_666217\|2020 | ···················· | ···················· | ···················· | ···················· | ···················· | ···················· |
| T | 2074 hCoV-19/Wales/PHWC-48B2C7/2020\|EPI_ISL_666218\|2020 | ···················· | ···················· | ···················· | ···················· | ···················· | ···················· |
| T | 2075 hCoV-19/Wales/PHWC-48B2D6/2020\|EPI_ISL_666219\|2020 | ···················· | ···················· | ···················· | ···················· | ···················· | ···················· |
| T | 2076 hCoV-19/Wales/PHWC-48B2E5/2020\|EPI_ISL_666220\|2020 | ···················· | ···················· | ···················· | ···················· | ···················· | ···················· |
| T | 2077 hCoV-19/Wales/PHWC-48B31F/2020\|EPI_ISL_666221\|2020 | ···················· | ···················· | ···················· | ···················· | ···················· | ···················· |
| T | 2078 hCoV-19/Wales/PHWC-48B32E/2020\|EPI_ISL_666222\|2020 | ···················· | ···················· | ···················· | ···················· | ···················· | ···················· |
| T | 2079 hCoV-19/Wales/PHWC-48B33D/2020\|EPI_ISL_666223\|2020 | ···················· | ···················· | ···················· | ···················· | ···················· | ···················· |
| T | 2080 hCoV-19/Wales/PHWC-48B34C/2020\|EPI_ISL_666224\|2020 | ···················· | ···················· | ···················· | ···················· | ···················· | ···················· |
| T | 2081 hCoV-19/Wales/PHWC-48B35B/2020\|EPI_ISL_666225\|2020 | ···················· | ···················· | ···················· | ···················· | ···················· | ···················· |
| T | 2082 hCoV-19/Wales/PHWC-48B36A/2020\|EPI_ISL_666226\|2020 | ···················· | ···················· | ···················· | ···················· | ···················· | ···················· |
| T | 2083 hCoV-19/Wales/PHWC-48B379/2020\|EPI_ISL_666227\|2020 | ···················· | ···················· | ···················· | ···················· | ···················· | ···················· |
| T | 2084 hCoV-19/Wales/PHWC-48B388/2020\|EPI_ISL_666228\|2020 | ···················· | ···················· | ···················· | ···················· | ···················· | ···················· |
| T | 2085 hCoV-19/Wales/PHWC-48B397/2020\|EPI_ISL_666229\|2020 | ···················· | ···················· | ···················· | ···················· | ···················· | ···················· |
| T | 2086 hCoV-19/Wales/PHWC-48B3A6/2020\|EPI_ISL_666230\|2020 | ···················· | ···················· | ···················· | ···················· | ···················· | ···················· |
| T | 2088 hCoV-19/Wales/PHWC-48B3D3/2020\|EPI_ISL_666232\|2020 | ···················· | ···················· | ···················· | ···················· | ···················· | ···················· |
| T | 2089 hCoV-19/Wales/PHWC-48B3E2/2020\|EPI_ISL_666233\|2020 | ···················· | ···················· | ···················· | ···················· | ···················· | ···················· |
| T | 2090 hCoV-19/Wales/PHWC-48B41C/2020\|EPI_ISL_666234\|2020 | ···················· | ···················· | ···················· | ···················· | ···················· | ···················· |
| T | 2091 hCoV-19/Wales/PHWC-48B42B/2020\|EPI_ISL_666235\|2020 | ···················· | ···················· | ···················· | ···················· | ···················· | ···················· |
| T | 2092 hCoV-19/Wales/PHWC-48B43A/2020\|EPI_ISL_666236\|2020 | ···················· | ···················· | ···················· | ···················· | ···················· | ···················· |
| T | 2093 hCoV-19/Wales/PHWC-48B449/2020\|EPI_ISL_666237\|2020 | ···················· | ···················· | ···················· | ···················· | ···················· | ···················· |
| T | 2094 hCoV-19/Wales/PHWC-48B467/2020\|EPI_ISL_666238\|2020 | ···················· | ···················· | ···················· | ···················· | ···················· | ···················· |
| T | 2095 hCoV-19/Wales/PHWC-48B476/2020\|EPI_ISL_666239\|2020 | ···················· | ···················· | ···················· | ···················· | ···················· | ···················· |
| T | 2096 hCoV-19/Wales/PHWC-48B494/2020\|EPI_ISL_666240\|2020 | ···················· | ···················· | ···················· | ···················· | ···················· | ···················· |
| T | 2097 hCoV-19/Wales/PHWC-48B4A3/2020\|EPI_ISL_666241\|2020 | ···················· | ···················· | ···················· | ···················· | ···················· | ···················· |
| T | 2098 hCoV-19/Wales/PHWC-48B4B2/2020\|EPI_ISL_666242\|2020 | ···················· | ···················· | ···················· | ···················· | ···················· | ···················· |
| T | 2099 hCoV-19/Wales/PHWC-48B4C1/2020\|EPI_ISL_666243\|2020 | ···················· | ···················· | ···················· | ···················· | ···················· | ···················· |
| T | 2100 hCoV-19/Wales/PHWC-48B4D0/2020\|EPI_ISL_666244\|2020 | ···················· | ···················· | ···················· | ···················· | ···················· | ···················· |
| T | 2101 hCoV-19/Wales/PHWC-48B4EF/2020\|EPI_ISL_666245\|2020 | ···················· | ···················· | ···················· | ···················· | ···················· | ···················· |
| T | 2102 hCoV-19/Wales/PHWC-48B4FE/2020\|EPI_ISL_666246\|2020 | ···················· | ···················· | ···················· | ···················· | ···················· | ···················· |
| T | 2103 hCoV-19/Wales/PHWC-48B50A/2020\|EPI_ISL_666247\|2020 | ···················· | ···················· | ···················· | ···················· | ···················· | ···················· |
| T | 2104 hCoV-19/Wales/PHWC-48B519/2020\|EPI_ISL_666248\|2020 | ···················· | ···················· | ···················· | ···················· | ···················· | ···················· |
| T | 2105 hCoV-19/Wales/PHWC-48B528/2020\|EPI_ISL_666249\|2020 | ···················· | ···················· | ···················· | ···················· | ···················· | ···················· |
| T | 2106 hCoV-19/Wales/PHWC-48B537/2020\|EPI_ISL_666250\|2020 | ···················· | ···················· | ···················· | ···················· | ···················· | ···················· |
| T | 2107 hCoV-19/Wales/PHWC-48B546/2020\|EPI_ISL_666251\|2020 | ···················· | ···················· | ···················· | ···················· | ···················· | ···················· |
| T | 2108 hCoV-19/Wales/PHWC-48B555/2020\|EPI_ISL_666252\|2020 | ···················· | ···················· | ···················· | ···················· | ···················· | ···················· |
| T | 2109 hCoV-19/Wales/PHWC-48B564/2020\|EPI_ISL_666253\|2020 | ···················· | ···················· | ···················· | ···················· | ···················· | ···················· |
| T | 2110 hCoV-19/Wales/PHWC-48B573/2020\|EPI_ISL_666254\|2020 | ···················· | ···················· | ···················· | ···················· | ···················· | ···················· |
| T | 2111 hCoV-19/Wales/PHWC-48B582/2020\|EPI_ISL_666255\|2020 | ···················· | ···················· | ···················· | ···················· | ···················· | ···················· |
| T | 2112 hCoV-19/Wales/PHWC-48B5BF/2020\|EPI_ISL_666256\|2020 | ···················· | ···················· | ···················· | ···················· | ···················· | ···················· |
| T | 2114 hCoV-19/Wales/PHWC-48B5DD/2020\|EPI_ISL_666258\|2020 | ···················· | ···················· | ···················· | ···················· | ···················· | ···················· |
| T | 2115 hCoV-19/Wales/PHWC-48B5EC/2020\|EPI_ISL_666259\|2020 | ···················· | ·A·················· | ···················· | ···················· | ···················· | ···A················ |
| T | 2116 hCoV-19/Wales/PHWC-48B607/2020\|EPI_ISL_666260\|2020 | ···················· | ···················· | ···················· | ···················· | ···················· | ···················· |
| T | 2117 hCoV-19/Wales/PHWC-48B616/2020\|EPI_ISL_666261\|2020 | ···················· | ···················· | ···················· | ···················· | ···················· | ···················· |
| T | 2118 hCoV-19/Wales/PHWC-48B625/2020\|EPI_ISL_666262\|2020 | ···················· | ···················· | ···················· | ···················· | ···················· | ···················· |
| T | 2119 hCoV-19/Wales/PHWC-48B634/2020\|EPI_ISL_666263\|2020 | ···················· | ···················· | ···················· | ···················· | ···················· | ···················· |
| T | 2120 hCoV-19/Wales/PHWC-48B643/2020\|EPI_ISL_666264\|2020 | ···················· | ···················· | ···················· | ···················· | ···················· | ···················· |
| T | 2121 hCoV-19/Wales/PHWC-48B652/2020\|EPI_ISL_666265\|2020 | ···················· | ···················· | ···················· | ···················· | ···················· | ···················· |

| | | | | | | | | |
|---|---|---|---|---|---|---|---|---|
| T | 2122 hCoV-19/Wales/PHWC-48B661/2020\|EPI_ISL_666266\|2020 | ···················· | ···················· | ···················· | ···················· | ···················· | ···················· | |
| T | 2123 hCoV-19/Wales/PHWC-48B69E/2020\|EPI_ISL_666267\|2020 | ···················· | ···················· | ···················· | ···················· | ···················· | ···················· | |
| T | 2124 hCoV-19/Wales/PHWC-48B6AD/2020\|EPI_ISL_666268\|2020 | ···················· | ···················· | ···················· | ···················· | ···················· | ···················· | |
| T | 2125 hCoV-19/Wales/PHWC-48B6DA/2020\|EPI_ISL_666269\|2020 | ···················· | ···················· | ···················· | ···················· | ···················· | ···················· | |
| T | 2126 hCoV-19/Wales/PHWC-48B6E9/2020\|EPI_ISL_666270\|2020 | ···················· | ···················· | ···················· | ···················· | ···················· | ···················· | |
| T | 2127 hCoV-19/Wales/PHWC-48B6F8/2020\|EPI_ISL_666271\|2020 | ···················· | ···················· | ···················· | ···················· | ···················· | ···················· | |
| T | 2128 hCoV-19/Wales/PHWC-48B704/2020\|EPI_ISL_666272\|2020 | ···················· | ···················· | ···················· | ···················· | ···················· | ···················· | |
| T | 2129 hCoV-19/Wales/PHWC-48B713/2020\|EPI_ISL_666273\|2020 | ···················· | ···················· | ···················· | ···················· | ···················· | ···················· | |
| T | 2130 hCoV-19/Wales/PHWC-48B722/2020\|EPI_ISL_666274\|2020 | ···················· | ···················· | ···················· | ···················· | ···················· | ···················· | |
| T | 2131 hCoV-19/Wales/PHWC-48B731/2020\|EPI_ISL_666275\|2020 | ···················· | ···················· | ···················· | ···················· | ···················· | ···················· | |
| T | 2132 hCoV-19/Wales/PHWC-48B740/2020\|EPI_ISL_666276\|2020 | ···················· | ···················· | ···················· | ···················· | ···················· | ···················· | |
| T | 2133 hCoV-19/Wales/PHWC-48B75F/2020\|EPI_ISL_666277\|2020 | ···················· | ···················· | ···················· | ···················· | ···················· | ···················· | |
| T | 2134 hCoV-19/Wales/PHWC-48B76E/2020\|EPI_ISL_666278\|2020 | ···················· | ···················· | ···················· | ···················· | ···················· | ····N··············· | |
| T | 2135 hCoV-19/Wales/PHWC-48B77D/2020\|EPI_ISL_666279\|2020 | ···················· | ···················· | ···················· | ···················· | ···················· | ···················· | |
| T | 2136 hCoV-19/Wales/PHWC-48B78C/2020\|EPI_ISL_666280\|2020 | ···················· | ···················· | ···················· | ···················· | ···················· | ···················· | |
| T | 2137 hCoV-19/Wales/PHWC-48B79B/2020\|EPI_ISL_666281\|2020 | ···················· | ···················· | ···················· | ···················· | NNN················· | ···················· | 1 |
| T | 2138 hCoV-19/Wales/PHWC-48B7AA/2020\|EPI_ISL_666282\|2020 | ···················· | ···················· | ···················· | ···················· | ···················· | ···················· | |
| T | 2139 hCoV-19/Wales/PHWC-48B7B9/2020\|EPI_ISL_666283\|2020 | ···················· | ···················· | ···················· | ···················· | ···················· | ···················· | |
| T | 2140 hCoV-19/Wales/PHWC-48B7C8/2020\|EPI_ISL_666284\|2020 | ···················· | ···················· | ···················· | ···················· | ···················· | ···················· | |
| T | 2141 hCoV-19/Wales/PHWC-48B7D7/2020\|EPI_ISL_666285\|2020 | ···················· | ···················· | ···················· | ···················· | ···················· | ···················· | |
| T | 2142 hCoV-19/Wales/PHWC-48B7F5/2020\|EPI_ISL_666286\|2020 | ···················· | ···················· | ···················· | ···················· | ···················· | ···················· | |
| T | 2143 hCoV-19/Wales/PHWC-48B810/2020\|EPI_ISL_666287\|2020 | ···················· | ···················· | ···················· | ···················· | ···················· | ···················· | |
| T | 2144 hCoV-19/Wales/PHWC-48B82F/2020\|EPI_ISL_666288\|2020 | ···················· | ···················· | ···················· | ···················· | ···················· | ···················· | |
| T | 2145 hCoV-19/Wales/PHWC-48B84D/2020\|EPI_ISL_666289\|2020 | ···················· | ···················· | ···················· | ···················· | ···················· | ···················· | |
| T | 2146 hCoV-19/Wales/PHWC-48B85C/2020\|EPI_ISL_666290\|2020 | ···················· | ···················· | ···················· | ···················· | ···················· | ···················· | |
| T | 2147 hCoV-19/Wales/PHWC-48B86B/2020\|EPI_ISL_666291\|2020 | ···················· | ···················· | ···················· | ···················· | ···················· | ···················· | |
| T | 2148 hCoV-19/Wales/PHWC-48B87A/2020\|EPI_ISL_666292\|2020 | ···················· | ···················· | ···················· | ···················· | ···················· | ···················· | |
| T | 2149 hCoV-19/Wales/PHWC-48B889/2020\|EPI_ISL_666293\|2020 | ···················· | ···················· | ···················· | ···················· | ···················· | ···················· | |
| T | 2150 hCoV-19/Wales/PHWC-48B898/2020\|EPI_ISL_666294\|2020 | ···················· | ···················· | ···················· | ···················· | ···················· | ···················· | |
| T | 2151 hCoV-19/Wales/PHWC-48B8A7/2020\|EPI_ISL_666295\|2020 | ···················· | ···················· | ···················· | ···················· | ···················· | ···················· | |
| T | 2152 hCoV-19/Wales/PHWC-48B8C5/2020\|EPI_ISL_666296\|2020 | ···················· | ···················· | ···················· | ···················· | ···················· | ···················· | |
| T | 2153 hCoV-19/Wales/PHWC-48B8D4/2020\|EPI_ISL_666297\|2020 | ···················· | ···················· | ···················· | ···················· | ···················· | ···················· | |
| T | 2154 hCoV-19/Wales/PHWC-48B8E3/2020\|EPI_ISL_666298\|2020 | ···················· | ···················· | ···················· | ···················· | ···················· | ···················· | |
| T | 2155 hCoV-19/Wales/PHWC-48B90E/2020\|EPI_ISL_666299\|2020 | ···················· | ···················· | ···················· | ···················· | ···················· | ···················· | |
| T | 2156 hCoV-19/USA/OR-OHSU-3179/2020\|EPI_ISL_667500\|2020- | ···················· | ···················· | ···················· | ···················· | ···················· | ···················· | |
| T | 2157 hCoV-19/USA/OR-OHSU-3183/2020\|EPI_ISL_667501\|2020- | ···················· | ···················· | ···················· | ···················· | ···················· | ···················· | |
| T | 2158 hCoV-19/USA/OR-OHSU-3545/2020\|EPI_ISL_667505\|2020- | ···················· | ·A·················· | ···················· | ···················· | ···················· | ···················· | |
| T | 2159 hCoV-19/USA/OR-OHSU-3561/2020\|EPI_ISL_667506\|2020- | ···················· | ···················· | ···················· | ···················· | ···················· | ···················· | |
| T | 2160 hCoV-19/USA/OR-OHSU-3577/2020\|EPI_ISL_667507\|2020- | ···················· | ···················· | ···················· | ···················· | ···················· | ···················· | |
| T | 2161 hCoV-19/USA/OR-OHSU-3680/2020\|EPI_ISL_667508\|2020- | ···················· | ·A·················· | ···················· | ···················· | ···················· | ···················· | |
| T | 2162 hCoV-19/USA/OR-OHSU-3692/2020\|EPI_ISL_667509\|2020- | ···················· | ···················· | ···················· | ···················· | ···················· | ···················· | |
| T | 2163 hCoV-19/USA/OR-OHSU-3702/2020\|EPI_ISL_667510\|2020- | ···················· | ···················· | ···················· | ···················· | ···················· | ···················· | |
| T | 2164 hCoV-19/USA/OR-OHSU-3743/2020\|EPI_ISL_667511\|2020- | ···················· | ···················· | ···················· | ···················· | ···················· | ···················· | |
| T | 2165 hCoV-19/USA/OR-OHSU-3777/2020\|EPI_ISL_667512\|2020- | ···················· | ···················· | ···················· | ···················· | ···················· | ···················· | |
| T | 2166 hCoV-19/USA/OR-OHSU-3845/2020\|EPI_ISL_667513\|2020- | ···················· | ·A·················· | ···················· | ···················· | ···················· | ···················· | |
| T | 2167 hCoV-19/USA/OR-OHSU-3865/2020\|EPI_ISL_667514\|2020- | ···················· | ·A·················· | ···················· | ···················· | ···················· | ···················· | |
| T | 2168 hCoV-19/USA/OR-OHSU-3875/2020\|EPI_ISL_667515\|2020- | ···················· | ···················· | ···················· | ···················· | ···················· | ···················· | |
| T | 2169 hCoV-19/USA/OR-OHSU-3912/2020\|EPI_ISL_667516\|2020- | ···················· | ·A·················· | ···················· | ···················· | ···················· | ···················· | |
| T | 2170 hCoV-19/USA/OR-OHSU-3995/2020\|EPI_ISL_667519\|2020- | ···················· | ···················· | ···················· | ···················· | ···················· | ···················· | |
| T | 2171 hCoV-19/USA/OR-OHSU-3996/2020\|EPI_ISL_667520\|2020- | ···················· | ···················· | ···················· | ···················· | ···················· | ···················· | |
| T | 2172 hCoV-19/USA/OR-OHSU-3997/2020\|EPI_ISL_667521\|2020- | ···················· | ·A·················· | ···················· | ···················· | ···················· | ···················· | |
| T | 2173 hCoV-19/USA/OR-OHSU-3999/2020\|EPI_ISL_667522\|2020- | ···················· | ···················· | ···················· | ···················· | ···················· | ···················· | |
| T | 2174 hCoV-19/USA/OR-OHSU-4032/2020\|EPI_ISL_667523\|2020- | ···················· | ·A·················· | ···················· | ···················· | ···················· | ···················· | |
| T | 2175 hCoV-19/USA/OR-OHSU-4034/2020\|EPI_ISL_667524\|2020- | ···················· | ···················· | ···················· | ···················· | ···················· | ···················· | |
| T | 2176 hCoV-19/USA/OR-OHSU-4068/2020\|EPI_ISL_667525\|2020- | ···················· | ···················· | ···················· | ···················· | ···················· | ···················· | |
| T | 2177 hCoV-19/USA/OR-OHSU-4117/2020\|EPI_ISL_667526\|2020- | ···················· | ·A·················· | ···················· | ···················· | ···················· | ···················· | |
| T | 2178 hCoV-19/USA/OR-OHSU-4128/2020\|EPI_ISL_667527\|2020- | ···················· | ···················· | ···················· | ···················· | ···················· | ···················· | |
| T | 2179 hCoV-19/USA/OR-OHSU-4137/2020\|EPI_ISL_667528\|2020- | ···················· | ···················· | ···················· | ···················· | ···················· | ···················· | |
| T | 2180 hCoV-19/USA/OR-OHSU-4152/2020\|EPI_ISL_667529\|2020- | ···················· | ···················· | ···················· | ···················· | ···················· | ···················· | |
| T | 2181 hCoV-19/USA/OR-OHSU-4154/2020\|EPI_ISL_667530\|2020- | ···················· | ·A·················· | ···················· | ···················· | ···················· | ···················· | |
| T | 2182 hCoV-19/USA/OR-OHSU-4155/2020\|EPI_ISL_667531\|2020- | ···················· | ·A·················· | ···················· | ···················· | ···················· | ···················· | |
| T | 2183 hCoV-19/USA/OR-OHSU-4156/2020\|EPI_ISL_667532\|2020- | ···················· | ···················· | ···················· | ···················· | ···················· | ···················· | |
| T | 2184 hCoV-19/USA/OR-OHSU-4157/2020\|EPI_ISL_667533\|2020- | ···················· | ···················· | ···················· | ···················· | ···················· | ···················· | |
| T | 2185 hCoV-19/USA/OR-OHSU-4292/2020\|EPI_ISL_667534\|2020- | ···················· | ···················· | ···················· | ···················· | ···················· | ···················· | |
| T | 2186 hCoV-19/USA/OR-OHSU-4315/2020\|EPI_ISL_667535\|2020- | ···················· | ···················· | ···················· | ···················· | ···················· | ···················· | |
| T | 2187 hCoV-19/USA/OR-OHSU-4346/2020\|EPI_ISL_667536\|2020- | ···················· | ···················· | ···················· | ···················· | ···················· | ···················· | |
| T | 2188 hCoV-19/USA/OR-OHSU-4446/2020\|EPI_ISL_667537\|2020- | ···················· | ···················· | ···················· | ···················· | ···················· | ···················· | |
| T | 2189 hCoV-19/USA/OR-OHSU-4584/2020\|EPI_ISL_667538\|2020- | ···················· | ···················· | ···················· | ···················· | ···················· | ···················· | |
| T | 2190 hCoV-19/USA/OR-OHSU-4348/2020\|EPI_ISL_667539\|2020- | ···················· | ···················· | ···················· | ···················· | ···················· | ···················· | |
| T | 2191 hCoV-19/USA/OR-OHSU-4349/2020\|EPI_ISL_667540\|2020- | ···················· | ·A·················· | ···················· | ···················· | ···················· | ···················· | |
| T | 2192 hCoV-19/USA/OR-OHSU-4387/2020\|EPI_ISL_667541\|2020- | ···················· | ···················· | ···················· | ···················· | ···················· | ···················· | |
| T | 2193 hCoV-19/USA/OR-OHSU-4397/2020\|EPI_ISL_667542\|2020- | ···················· | ···················· | ···················· | ···················· | ···················· | ···················· | |
| T | 2194 hCoV-19/USA/OR-OHSU-4500/2020\|EPI_ISL_667543\|2020- | ···················· | ···················· | ···················· | ···················· | ···················· | ···················· | |
| T | 2195 hCoV-19/USA/OR-OHSU-4663/2020\|EPI_ISL_667544\|2020- | ···················· | ···················· | ···················· | ···················· | ···················· | ···················· | |
| T | 2196 hCoV-19/USA/OR-OHSU-4667/2020\|EPI_ISL_667545\|2020- | ···················· | ···················· | ···················· | ···················· | ···················· | ···················· | |
| T | 2197 hCoV-19/USA/OR-OHSU-4675/2020\|EPI_ISL_667546\|2020- | ···················· | ···················· | ···················· | ···················· | ···················· | ···················· | |
| T | 2198 hCoV-19/USA/OR-OHSU-4702/2020\|EPI_ISL_667547\|2020- | ···················· | ···················· | ···················· | ···················· | ···················· | ···················· | |
| T | 2199 hCoV-19/USA/OR-OHSU-4788/2020\|EPI_ISL_667548\|2020- | ···················· | ·A·················· | ···················· | ···················· | ···················· | ···················· | |
| T | 2200 hCoV-19/USA/OR-OHSU-5184/2020\|EPI_ISL_667549\|2020- | ···················· | ·············A······ | ···················· | ···················· | ···················· | ···················· | |
| T | 2201 hCoV-19/USA/OR-OHSU-2047/2020\|EPI_ISL_667551\|2020- | ···················· | ···················· | ···················· | ···················· | ···················· | ···················· | |

T 2202 hCoV-19/USA/OR-OHSU-2063/2020|EPI_ISL_667552|2020- ···················· ···················· ····················· ···················· ···················· ····················
T 2203 hCoV-19/USA/OR-OHSU-2157/2020|EPI_ISL_667553|2020- ···················· ···················· ····················· ···················· ···················· ····················
T 2204 hCoV-19/Japan/IC-0130/2020|EPI_ISL_667559|2020-08 ···················· ···················· ····················· ···················· ···················· ····················
T 2205 hCoV-19/Japan/IC-0131/2020|EPI_ISL_667560|2020-09 ···················· ···················· ····················· ···················· ···················· ····················
T 2206 hCoV-19/Japan/IC-0132/2020|EPI_ISL_667561|2020-07 ···················· ···················· ····················· ···················· ···················· ····················
T 2207 hCoV-19/Japan/IC-0133/2020|EPI_ISL_667562|2020-07 ···················· ···················· ····················· ···················· ···················· ····················
T 2208 hCoV-19/Japan/IC-0134/2020|EPI_ISL_667563|2020-07 ···················· ···················· ····················· ···················· ···················· ····················
T 2209 hCoV-19/Japan/IC-0135/2020|EPI_ISL_667564|2020-07 ···················· ···················· ····················· ···················· ···················· ····················
T 2210 hCoV-19/Japan/IC-0137/2020|EPI_ISL_667566|2020-09 ···················· ···················· ····················· ···················· ···················· ····················
T 2211 hCoV-19/Japan/IC-0138/2020|EPI_ISL_667567|2020-08 ···················· ···················· ····················· ···················· ···················· ····················
T 2212 hCoV-19/Japan/IC-0139/2020|EPI_ISL_667568|2020-09 ···················· ···················· ····················· ···················· ···················· ····················
T 2213 hCoV-19/Japan/IC-0140/2020|EPI_ISL_667569|2020-09 ···················· ···················· ····················· ···················· ···················· ····················
T 2214 hCoV-19/Japan/IC-0141/2020|EPI_ISL_667570|2020-09 ···················· ·········A·········· ····················· ···················· ···················· ····················
T 2215 hCoV-19/Japan/IC-0142/2020|EPI_ISL_667571|2020-08 ···················· ···················· ····················· ···················· ···················· ····················
T 2216 hCoV-19/Japan/IC-0143/2020|EPI_ISL_667572|2020-08 ···················· ···················· ····················· ···················· ···················· ····················
T 2217 hCoV-19/Japan/IC-0144/2020|EPI_ISL_667573|2020-09 ···················· ···················· ····················· ···················· ···················· ····················
T 2218 hCoV-19/Japan/IC-0145/2020|EPI_ISL_667574|2020-09 ···················· ···················· ····················· ···················· ···················· ····················
T 2219 hCoV-19/Japan/IC-0146/2020|EPI_ISL_667575|2020-09 ···················· ·A·················· ····················· ···················· ···················· ····················
T 2220 hCoV-19/Japan/IC-0147/2020|EPI_ISL_667576|2020-07 ···················· ···················· ····················· ···················· ···················· ····················
T 2221 hCoV-19/Japan/IC-0148/2020|EPI_ISL_667577|2020-08 ···················· ···················· ····················· ···················· ···················· ····················
T 2222 hCoV-19/Japan/IC-0149/2020|EPI_ISL_667578|2020-08 ···················· ···················· ····················· ···················· ···················· ····················
T 2223 hCoV-19/Japan/IC-0150/2020|EPI_ISL_667579|2020-08 ···················· ···················· ····················· ···················· ···················· ····················
T 2224 hCoV-19/Japan/IC-0151/2020|EPI_ISL_667580|2020-08 ···················· ···················· ····················· ···················· ···················· ····················
T 2225 hCoV-19/Japan/IC-0152/2020|EPI_ISL_667581|2020-08 ···················· ···················· ····················· ···················· ···················· ····················
T 2226 hCoV-19/Japan/IC-0153/2020|EPI_ISL_667582|2020-08 ···················· ···················· ····················· ···················· ···················· ····················
T 2227 hCoV-19/Japan/IC-0154/2020|EPI_ISL_667583|2020-08 ···················· ···················· ····················· ···················· ···················· ····················
T 2228 hCoV-19/Japan/IC-0155/2020|EPI_ISL_667584|2020-08 ···················· ···················· ····················· ···················· ···················· ····················
T 2229 hCoV-19/Japan/IC-0156/2020|EPI_ISL_667585|2020-08 ···················· ···················· ····················· ···················· ···················· ····················
T 2230 hCoV-19/Japan/IC-0157/2020|EPI_ISL_667586|2020-08 ···················· ······A············· ····················· ···················· ···················· ····················
T 2231 hCoV-19/Japan/IC-0158/2020|EPI_ISL_667587|2020-09 ···················· ·A·················· ····················· ···················· ···················· ···A················
T 2232 hCoV-19/Japan/IC-0159/2020|EPI_ISL_667588|2020-09 ···················· ·A·················· ····················· ···················· ···················· ···A················
T 2233 hCoV-19/Japan/IC-0160/2020|EPI_ISL_667589|2020-09 ···················· ···················· ····················· ···················· ···················· ····················
T 2234 hCoV-19/Japan/IC-0161/2020|EPI_ISL_667590|2020-09 ···················· ···················· ····················· ···················· ···················· ····················
T 2235 hCoV-19/Japan/IC-0162/2020|EPI_ISL_667591|2020-09 ···················· ···················· ····················· ···················· ···················· ····················
T 2236 hCoV-19/Japan/IC-0163/2020|EPI_ISL_667592|2020-09 ···················· ·A·················· ····················· ···················· ···················· ···A················
T 2237 hCoV-19/Japan/IC-0165/2020|EPI_ISL_667594|2020-09 ···················· ···················· ····················· ···················· ···················· ····················
T 2238 hCoV-19/Japan/IC-0166/2020|EPI_ISL_667595|2020-09 ···················· ···················· ····················· ···················· ···················· ····················
T 2239 hCoV-19/Japan/IC-0167/2020|EPI_ISL_667596|2020-09 ···················· ···················· ····················· ···················· ···················· ····················
T 2240 hCoV-19/Japan/IC-0168/2020|EPI_ISL_667597|2020-09 ···················· ·A·················· ····················· ···················· ···················· ····················
T 2241 hCoV-19/Japan/IC-0169/2020|EPI_ISL_667598|2020-09 ···················· ···················· ····················· ···················· ···················· ····················
T 2242 hCoV-19/Japan/IC-0170/2020|EPI_ISL_667599|2020-09 ···················· ···················· ····················· ···················· ···················· ····················
T 2243 hCoV-19/Denmark/DCGC-10012/2020|EPI_ISL_668837|202 ···················· ···················· ····················· ···················· ···················· ····················
T 2244 hCoV-19/Denmark/DCGC-9213/2020|EPI_ISL_668853|2020 ···················· ···················· ····················· ···················· ···················· ····················
T 2245 hCoV-19/Denmark/DCGC-9214/2020|EPI_ISL_668854|2020 ···················· ······N············· ····················· ···················· ···················· ····················
T 2246 hCoV-19/Denmark/DCGC-9215/2020|EPI_ISL_668855|2020 ···················· ···················· ····················· ···················· ···················· ····················
T 2247 hCoV-19/Denmark/DCGC-9219/2020|EPI_ISL_668856|2020 ···················· ·A·················· ····················· ···················· ···················· ···A················
T 2248 hCoV-19/Denmark/DCGC-9220/2020|EPI_ISL_668857|2020 ···················· ···················· ····················· ···················· ···················· ····················
T 2249 hCoV-19/Denmark/DCGC-9222/2020|EPI_ISL_668858|2020 ···················· ···················· ····················· ···················· ···················· ····················
T 2250 hCoV-19/Denmark/DCGC-9224/2020|EPI_ISL_668859|2020 ···················· ···················· ····················· ···················· ···················· ····················
T 2251 hCoV-19/Denmark/DCGC-9225/2020|EPI_ISL_668860|2020 ···················· ···················· ····················· ···················· ···················· ····················
T 2252 hCoV-19/Denmark/DCGC-9226/2020|EPI_ISL_668861|2020 ···················· ···················· ····················· ···················· ···················· ····················
T 2253 hCoV-19/Denmark/DCGC-9227/2020|EPI_ISL_668862|2020 ···················· ···················· ····················· ···················· ···················· ····················
T 2254 hCoV-19/Denmark/DCGC-9234/2020|EPI_ISL_668863|2020 ···················· ···················· ····················· ···················· ···················· ····················
T 2255 hCoV-19/Denmark/DCGC-9236/2020|EPI_ISL_668864|2020 ···················· ···················· ····················· ···················· ···················· ····················
T 2256 hCoV-19/Denmark/DCGC-9238/2020|EPI_ISL_668865|2020 ···················· ···················· ····················· ···················· ···················· ····················
T 2257 hCoV-19/Denmark/DCGC-9239/2020|EPI_ISL_668866|2020 ···················· ···················· ····················· ···················· ···················· ····················
T 2258 hCoV-19/Denmark/DCGC-9240/2020|EPI_ISL_668867|2020 ···················· ·A·················· ····················· ···················· ···················· ····················
T 2259 hCoV-19/Denmark/DCGC-9242/2020|EPI_ISL_668868|2020 ···················· ···················· ····················· ···················· ···················· ····················
T 2260 hCoV-19/Denmark/DCGC-9243/2020|EPI_ISL_668869|2020 ···················· ·A·················· ····················· ···················· ···················· ····················
T 2261 hCoV-19/Denmark/DCGC-9244/2020|EPI_ISL_668870|2020 ···················· ···················· ····················· ···················· ···················· ····················
T 2262 hCoV-19/Denmark/DCGC-9245/2020|EPI_ISL_668871|2020 ···················· ···················· ····················· ···················· ···················· ····················
T 2263 hCoV-19/Denmark/DCGC-9247/2020|EPI_ISL_668872|2020 ···················· ·A·················· ····················· ···················· ···················· ····················
T 2264 hCoV-19/Denmark/DCGC-9248/2020|EPI_ISL_668873|2020 ···················· ···················· ····················· ···················· ···················· ····················
T 2265 hCoV-19/Denmark/DCGC-9251/2020|EPI_ISL_668874|2020 ···················· ·A·················· ····················· ···················· ···················· ····················
T 2266 hCoV-19/Denmark/DCGC-9253/2020|EPI_ISL_668875|2020 ···················· ···················· ····················· ···················· ······T············· ····················
T 2267 hCoV-19/Denmark/DCGC-9257/2020|EPI_ISL_668876|2020 ···················· ···················· ····················· ···················· ···················· ····················
T 2268 hCoV-19/Denmark/DCGC-9258/2020|EPI_ISL_668877|2020 ···················· ···················· ····················· ···················· ···················· ····················
T 2269 hCoV-19/Denmark/DCGC-9260/2020|EPI_ISL_668878|2020 ···················· ···················· ····················· ···················· ···················· ····················
T 2270 hCoV-19/Denmark/DCGC-9264/2020|EPI_ISL_668879|2020 ···················· ···················· ····················· ···················· ···················· ····················
T 2271 hCoV-19/Denmark/DCGC-9270/2020|EPI_ISL_668880|2020 ···················· ···················· ····················· ···················· ···················· ····················
T 2272 hCoV-19/Denmark/DCGC-9273/2020|EPI_ISL_668881|2020 ···················· ·A·················· ····················· ···················· ···················· ····················
T 2273 hCoV-19/Denmark/DCGC-9275/2020|EPI_ISL_668882|2020 ···················· ···················· ····················· ···················· ···················· ····················
T 2274 hCoV-19/Denmark/DCGC-9276/2020|EPI_ISL_668883|2020 ···················· ···················· ····················· ···················· ···················· ····················
T 2275 hCoV-19/Denmark/DCGC-9278/2020|EPI_ISL_668884|2020 ···················· ···················· ····················· ···················· ···················· ····················
T 2276 hCoV-19/Denmark/DCGC-9279/2020|EPI_ISL_668885|2020 ···················· ···················· ····················· ···················· ···················· ····················
T 2277 hCoV-19/Denmark/DCGC-9282/2020|EPI_ISL_668886|2020 ···················· ···················· ····················· ···················· ···················· ····················
T 2278 hCoV-19/Denmark/DCGC-9283/2020|EPI_ISL_668887|2020 ···················· ···················· ····················· ···················· ···················· ····················
T 2279 hCoV-19/Denmark/DCGC-9284/2020|EPI_ISL_668888|2020 ···················· ···················· ····················· ···················· ···················· ····················
T 2280 hCoV-19/Denmark/DCGC-9291/2020|EPI_ISL_668889|2020 ···················· ···················· ····················· ···················· ···················· ····················
T 2281 hCoV-19/Denmark/DCGC-9293/2020|EPI_ISL_668890|2020 ···················· ···················· ····················· ···················· ···················· ····················

| | | | | | | | |
|---|---|---|---|---|---|---|---|
| T | 2282 hCoV-19/Denmark/DCGC-9296/2020\|EPI_ISL_668891\|2020 | ···················· | ···················· | ···················· | ···················· | ·······T············ | ···················· |
| T | 2283 hCoV-19/Denmark/DCGC-9297/2020\|EPI_ISL_668892\|2020 | ···················· | ···················· | ···················· | ···················· | ···················· | ···················· |
| T | 2284 hCoV-19/Denmark/DCGC-9298/2020\|EPI_ISL_668893\|2020 | ···················· | ···················· | ···················· | ···················· | ···················· | ···················· |
| T | 2285 hCoV-19/Denmark/DCGC-9299/2020\|EPI_ISL_668894\|2020 | ···················· | ···················· | ···················· | ···················· | ···················· | ···················· |
| T | 2286 hCoV-19/Denmark/DCGC-9300/2020\|EPI_ISL_668895\|2020 | ···················· | ···················· | ···················· | ···················· | ···················· | ···················· |
| T | 2287 hCoV-19/Denmark/DCGC-9302/2020\|EPI_ISL_668896\|2020 | ···················· | ···················· | ···················· | ···················· | ···················· | ···················· |
| T | 2288 hCoV-19/Denmark/DCGC-9303/2020\|EPI_ISL_668897\|2020 | ···················· | ···················· | ···················· | ···················· | ·······T············ | ···················· |
| T | 2289 hCoV-19/Denmark/DCGC-9304/2020\|EPI_ISL_668898\|2020 | ···················· | ···················· | ···················· | ···················· | ···················· | ···················· |
| T | 2290 hCoV-19/Denmark/DCGC-9305/2020\|EPI_ISL_668899\|2020 | ···················· | ···················· | ···················· | ···················· | ···················· | ···················· |
| T | 2291 hCoV-19/Denmark/DCGC-9935/2020\|EPI_ISL_669280\|2020 | ···················· | ···················· | ···················· | ···················· | ···················· | ···················· |
| T | 2292 hCoV-19/Denmark/DCGC-10888/2020\|EPI_ISL_670266\|202 | ···················· | ···················· | ···················· | ···················· | ···················· | ···················· |
| T | 2293 hCoV-19/Denmark/DCGC-10893/2020\|EPI_ISL_670269\|202 | ···················· | ···················· | ···················· | ···················· | ···················· | ···················· |
| T | 2294 hCoV-19/Canada/ON-PHL-20-02457/2020\|EPI_ISL_671598 | ···················· | ···················· | ···················· | ···················· | ···················· | ···················· |
| T | 2295 hCoV-19/Canada/ON-PHL-20-02456/2020\|EPI_ISL_671599 | ···················· | ···················· | ···················· | ···················· | ···················· | ···················· |
| T | 2296 hCoV-19/USA/CA-CZB-13274/2020\|EPI_ISL_672085\|2020- | ···················· | ·A·················· | ···················· | ···················· | ···················· | ···················· |
| T | 2297 hCoV-19/USA/CA-CZB-13385/2020\|EPI_ISL_672086\|2020- | ···················· | ·A·················· | ···················· | ···················· | ···················· | ···················· |
| T | 2298 hCoV-19/USA/CA-CZB-5745/2020\|EPI_ISL_672087\|2020-0 | ···················· | ·A·················· | ···················· | ···················· | ···················· | ···················· |
| T | 2299 hCoV-19/USA/CA-CZB-12978/2020\|EPI_ISL_672088\|2020- | ···················· | ···················· | ···················· | ···················· | ···················· | ···················· |
| T | 2300 hCoV-19/USA/CA-CZB-11883/2020\|EPI_ISL_672089\|2020- | ···················· | ·A·················· | ···················· | ···················· | ···················· | ···················· |
| T | 2301 hCoV-19/USA/CA-CZB-13501/2020\|EPI_ISL_672098\|2020- | ···················· | ·A·················· | ···················· | ···················· | ···················· | ···················· |
| T | 2303 hCoV-19/USA/CA-CZB-13491/2020\|EPI_ISL_672100\|2020- | ···················· | ·A·················· | ···················· | ···················· | ···················· | ···················· |
| T | 2304 hCoV-19/USA/CA-CZB-13471/2020\|EPI_ISL_672101\|2020- | ···················· | ·A·················· | ···················· | ···················· | ···················· | ···················· |
| T | 2305 hCoV-19/England/QEUH-AF4C6B/2020\|EPI_ISL_663101\|20 | ···················· | ···················· | ···················· | ···················· | ···················· | ···················· |
| T | 2306 hCoV-19/England/QEUH-AF4BB9/2020\|EPI_ISL_663102\|20 | ···················· | ···················· | ···················· | ···················· | ···················· | ···················· |
| T | 2307 hCoV-19/England/QEUH-AF5796/2020\|EPI_ISL_663103\|20 | ···················· | ···················· | ···················· | ···················· | ···················· | ···················· |
| T | 2308 hCoV-19/Wales/PHWC-48AD03/2020\|EPI_ISL_665750\|2020 | ···················· | ···················· | ···················· | ···················· | ···················· | ···················· |
| T | 2309 hCoV-19/Wales/PHWC-48B0CD/2020\|EPI_ISL_665751\|2020 | ···················· | ···················· | ···················· | ···················· | ···················· | ···················· |
| T | 2310 hCoV-19/Wales/PHWC-48B2F4/2020\|EPI_ISL_665752\|2020 | ···················· | ···················· | ···················· | ···················· | ···················· | ···················· |
| T | 2311 hCoV-19/Wales/PHWC-48B3B5/2020\|EPI_ISL_665753\|2020 | ···················· | ···················· | ···················· | ···················· | ···················· | ···················· |
| T | 2312 hCoV-19/Wales/PHWC-48BDF3/2020\|EPI_ISL_665754\|2020 | ···················· | ···················· | ···················· | ···················· | ···················· | ···················· |
| T | 2313 hCoV-19/Wales/PHWC-48C202/2020\|EPI_ISL_665755\|2020 | ···················· | ···················· | ···················· | ···················· | ···················· | ···················· |
| T | 2314 hCoV-19/Wales/PHWC-48C2A8/2020\|EPI_ISL_665756\|2020 | ···················· | ···················· | ···················· | ···················· | ···················· | ···················· |
| T | 2315 hCoV-19/England/LOND-1251144/2020\|EPI_ISL_665757\|2 | ···················· | ···················· | ···················· | ···················· | ···················· | ···················· |
| T | 2316 hCoV-19/England/LOND-1251162/2020\|EPI_ISL_665758\|2 | ···················· | ···················· | ···················· | ···················· | ···················· | ···················· |
| T | 2317 hCoV-19/England/LOND-1250FC3/2020\|EPI_ISL_665759\|2 | ···················· | ···················· | ···················· | ···················· | ···················· | ···················· |
| T | 2319 hCoV-19/England/QEUH-A9C9AA/2020\|EPI_ISL_665761\|20 | ···················· | ···················· | ···················· | ···················· | ···················· | ···················· |
| T | 2320 hCoV-19/Scotland/EDB9367/2020\|EPI_ISL_665762\|2020- | ···················· | ···················· | ···················· | ···················· | ···················· | ···················· |
| T | 2321 hCoV-19/Wales/PHWC-48A9E1/2020\|EPI_ISL_665763\|2020 | ···················· | ···················· | ···················· | ···················· | ···················· | ···················· |
| T | 2322 hCoV-19/Wales/PHWC-48AD5E/2020\|EPI_ISL_665764\|2020 | ···················· | ···················· | ···················· | ···················· | ···················· | ···················· |
| T | 2323 hCoV-19/Wales/PHWC-48AD7C/2020\|EPI_ISL_665765\|2020 | ···················· | ···················· | ···················· | ···················· | ···················· | ···················· |
| T | 2324 hCoV-19/Denmark/DCGC-9831/2020\|EPI_ISL_669209\|2020 | ···················· | ···················· | ···················· | ···················· | ···················· | ···················· |
| T | 2325 hCoV-19/Denmark/DCGC-9832/2020\|EPI_ISL_669210\|2020 | ···················· | ···················· | ···················· | ···················· | ···················· | ···················· |
| T | 2326 hCoV-19/Denmark/DCGC-9833/2020\|EPI_ISL_669211\|2020 | ···················· | ···················· | ···················· | ···················· | ···················· | ···················· |
| T | 2327 hCoV-19/Denmark/DCGC-9834/2020\|EPI_ISL_669212\|2020 | ···················· | ···················· | ···················· | ···················· | ···················· | ···················· |
| T | 2328 hCoV-19/Denmark/DCGC-9835/2020\|EPI_ISL_669213\|2020 | ···················· | ···················· | ···················· | ···················· | ···················· | ···················· |
| T | 2329 hCoV-19/Denmark/DCGC-9836/2020\|EPI_ISL_669214\|2020 | ···················· | ···················· | ···················· | ···················· | ···················· | ···················· |
| T | 2330 hCoV-19/Denmark/DCGC-9837/2020\|EPI_ISL_669215\|2020 | ···················· | ···················· | ···················· | ···················· | ···················· | ···················· |
| T | 2331 hCoV-19/Denmark/DCGC-9838/2020\|EPI_ISL_669216\|2020 | ···················· | ···················· | ···················· | ···················· | ···················· | ···················· |
| T | 2332 hCoV-19/Denmark/DCGC-9839/2020\|EPI_ISL_669217\|2020 | ···················· | ···················· | ···················· | ···················· | ···················· | ···················· |
| T | 2333 hCoV-19/Denmark/DCGC-9840/2020\|EPI_ISL_669218\|2020 | ···················· | ···················· | ···················· | ···················· | ···················· | ···················· |
| T | 2334 hCoV-19/Denmark/DCGC-9841/2020\|EPI_ISL_669219\|2020 | ···················· | ·A·················· | ···················· | ···················· | ···················· | ···A················ |
| T | 2335 hCoV-19/Denmark/DCGC-9843/2020\|EPI_ISL_669220\|2020 | ···················· | ···················· | ···················· | ···················· | ···················· | ···················· |
| T | 2336 hCoV-19/Denmark/DCGC-9844/2020\|EPI_ISL_669221\|2020 | ···················· | ···················· | ···················· | ···················· | ···················· | ···················· |
| T | 2337 hCoV-19/Denmark/DCGC-9845/2020\|EPI_ISL_669222\|2020 | ···················· | ····A··············· | ···················· | ···················· | ···················· | ···················· |
| T | 2338 hCoV-19/Denmark/DCGC-9846/2020\|EPI_ISL_669223\|2020 | ···················· | ···················· | ···················· | ···················· | ···················· | ···················· |
| T | 2339 hCoV-19/Denmark/DCGC-9847/2020\|EPI_ISL_669224\|2020 | ···················· | ···················· | ···················· | ···················· | ···················· | ···················· |
| T | 2340 hCoV-19/Denmark/DCGC-9848/2020\|EPI_ISL_669225\|2020 | ···················· | ···················· | ···················· | ···················· | ···················· | ···················· |
| T | 2341 hCoV-19/Denmark/DCGC-9849/2020\|EPI_ISL_669226\|2020 | ···················· | ···················· | ···················· | ···················· | ···················· | ···················· |
| T | 2342 hCoV-19/Denmark/DCGC-9850/2020\|EPI_ISL_669227\|2020 | ···················· | ···················· | ···················· | ···················· | ···················· | ···················· |
| T | 2343 hCoV-19/Denmark/DCGC-9852/2020\|EPI_ISL_669228\|2020 | ···················· | ···················· | ···················· | ···················· | ···················· | ···················· |
| T | 2344 hCoV-19/Denmark/DCGC-9853/2020\|EPI_ISL_669229\|2020 | ···················· | ···················· | ···················· | ···················· | ···················· | ···················· |
| T | 2345 hCoV-19/Denmark/DCGC-9855/2020\|EPI_ISL_669230\|2020 | ···················· | ···················· | ···················· | ···················· | ···················· | ···················· |
| T | 2346 hCoV-19/Denmark/DCGC-9856/2020\|EPI_ISL_669231\|2020 | ···················· | ···················· | ···················· | ···················· | ···················· | ···················· |
| T | 2347 hCoV-19/Denmark/DCGC-9857/2020\|EPI_ISL_669232\|2020 | ···················· | ·A·················· | ···················· | ···················· | ···················· | ···A················ |
| T | 2348 hCoV-19/Denmark/DCGC-9858/2020\|EPI_ISL_669233\|2020 | ···················· | ···················· | ···················· | ···················· | ······T············· | ···················· |
| T | 2349 hCoV-19/Denmark/DCGC-9859/2020\|EPI_ISL_669234\|2020 | ···················· | ···················· | ···················· | ···················· | ···················· | ···················· |
| T | 2350 hCoV-19/Denmark/DCGC-9860/2020\|EPI_ISL_669235\|2020 | ···················· | ···················· | ···················· | ···················· | ···················· | ···················· |
| T | 2351 hCoV-19/Denmark/DCGC-9861/2020\|EPI_ISL_669236\|2020 | ···················· | ···················· | ···················· | ···················· | ···················· | ···················· |
| T | 2352 hCoV-19/Denmark/DCGC-9864/2020\|EPI_ISL_669237\|2020 | ···················· | ···················· | ···················· | ···················· | ···················· | ···················· |
| T | 2353 hCoV-19/Denmark/DCGC-9865/2020\|EPI_ISL_669238\|2020 | ···················· | ···················· | ···················· | ···················· | ···················· | ···················· |
| T | 2354 hCoV-19/Denmark/DCGC-9867/2020\|EPI_ISL_669239\|2020 | ···················· | ···················· | ···················· | ···················· | ···················· | ···················· |
| T | 2355 hCoV-19/Denmark/DCGC-9868/2020\|EPI_ISL_669240\|2020 | ···················· | ···················· | ···················· | ···················· | ···················· | ···················· |
| T | 2356 hCoV-19/Denmark/DCGC-9869/2020\|EPI_ISL_669241\|2020 | ···················· | ···················· | ···················· | ···················· | ···················· | ···················· |
| T | 2357 hCoV-19/Denmark/DCGC-9870/2020\|EPI_ISL_669242\|2020 | ···················· | ···················· | ···················· | ···················· | ···················· | ···················· |
| T | 2358 hCoV-19/Denmark/DCGC-9873/2020\|EPI_ISL_669243\|2020 | ···················· | ···················· | ···················· | ···················· | ···················· | ···················· |
| T | 2359 hCoV-19/Denmark/DCGC-9874/2020\|EPI_ISL_669244\|2020 | ···················· | ···················· | ···················· | ···················· | ···················· | ···················· |
| T | 2360 hCoV-19/Denmark/DCGC-9875/2020\|EPI_ISL_669245\|2020 | ···················· | ···················· | ···················· | ···················· | ···················· | ···················· |
| T | 2361 hCoV-19/Denmark/DCGC-9876/2020\|EPI_ISL_669246\|2020 | ···················· | ···················· | ···················· | ···················· | ···················· | ···················· |
| T | 2362 hCoV-19/Denmark/DCGC-9877/2020\|EPI_ISL_669247\|2020 | ···················· | ···················· | ···················· | ···················· | ···················· | ···················· |
| T | 2363 hCoV-19/Denmark/DCGC-9878/2020\|EPI_ISL_669248\|2020 | ···················· | ···················· | ···················· | ···················· | ···················· | ···················· |

| | | | | | | | |
|---|---|---|---|---|---|---|---|
| T | 2364 hCoV-19/Denmark/DCGC-9879/2020\|EPI_ISL_669249\|2020 | .................... | .................... | .................... | .................... | .................... | .................... |
| T | 2365 hCoV-19/Denmark/DCGC-9880/2020\|EPI_ISL_669250\|2020 | .................... | .................... | .................... | .................... | .................... | .................... |
| T | 2366 hCoV-19/Denmark/DCGC-9881/2020\|EPI_ISL_669251\|2020 | .................... | .................... | .................... | .................... | .................... | .................... |
| T | 2367 hCoV-19/Denmark/DCGC-9882/2020\|EPI_ISL_669252\|2020 | .................... | .................... | .................... | .................... | .................... | .................... |
| T | 2368 hCoV-19/Denmark/DCGC-9883/2020\|EPI_ISL_669253\|2020 | .................... | .................... | .................... | .................... | .................... | .................... |
| T | 2369 hCoV-19/Denmark/DCGC-9884/2020\|EPI_ISL_669254\|2020 | .................... | .................... | .................... | .................... | .................... | .................... |
| T | 2370 hCoV-19/Denmark/DCGC-9887/2020\|EPI_ISL_669255\|2020 | .................... | .................... | .................... | .................... | .................... | .................... |
| T | 2371 hCoV-19/Denmark/DCGC-9888/2020\|EPI_ISL_669256\|2020 | .................... | .................... | .................... | .................... | .................... | .................... |
| T | 2372 hCoV-19/Denmark/DCGC-9891/2020\|EPI_ISL_669257\|2020 | .................... | .A.................. | .................... | .................... | .................... | ..A................. |
| T | 2373 hCoV-19/Denmark/DCGC-9892/2020\|EPI_ISL_669258\|2020 | .................... | .................... | .................... | .................... | .................... | .................... |
| T | 2374 hCoV-19/Denmark/DCGC-9895/2020\|EPI_ISL_669259\|2020 | .................... | .A.................. | .................... | .................... | .................... | ..A................. |
| T | 2375 hCoV-19/Denmark/DCGC-9897/2020\|EPI_ISL_669260\|2020 | .................... | .................... | .................... | .................... | .................... | .................... |
| T | 2376 hCoV-19/Denmark/DCGC-9898/2020\|EPI_ISL_669261\|2020 | .................... | .................... | .................... | .................... | .................... | .................... |
| T | 2377 hCoV-19/Denmark/DCGC-9899/2020\|EPI_ISL_669262\|2020 | .................... | .................... | .................... | .................... | .................... | .................... |
| T | 2378 hCoV-19/Denmark/DCGC-9903/2020\|EPI_ISL_669263\|2020 | .................... | .A.................. | .................... | .................... | .................... | ..A................. |
| T | 2379 hCoV-19/Denmark/DCGC-9904/2020\|EPI_ISL_669264\|2020 | .................... | .................... | .................... | .................... | .................... | .................... |
| T | 2380 hCoV-19/Denmark/DCGC-9905/2020\|EPI_ISL_669265\|2020 | .................... | .................... | .................... | .................... | .................... | .................... |
| T | 2381 hCoV-19/Denmark/DCGC-9906/2020\|EPI_ISL_669266\|2020 | .................... | .................... | .................... | .................... | .................... | .................... |
| T | 2382 hCoV-19/Denmark/DCGC-9907/2020\|EPI_ISL_669267\|2020 | .................... | .................... | .................... | .................... | .................... | .................... |
| T | 2383 hCoV-19/Denmark/DCGC-9908/2020\|EPI_ISL_669268\|2020 | .................... | .A.................. | .................... | .................... | .................... | ..A................. |
| T | 2384 hCoV-19/Denmark/DCGC-9914/2020\|EPI_ISL_669269\|2020 | .................... | .................... | .................... | .................... | .................... | .................... |
| T | 2385 hCoV-19/Denmark/DCGC-9916/2020\|EPI_ISL_669270\|2020 | .................... | .................... | .................... | .................... | .................... | .................... |
| T | 2386 hCoV-19/Denmark/DCGC-9919/2020\|EPI_ISL_669271\|2020 | .................... | .................... | .................... | .................... | .................... | .................... |
| T | 2387 hCoV-19/Denmark/DCGC-9921/2020\|EPI_ISL_669272\|2020 | .................... | .................... | .................... | .................... | .................... | .................... |
| T | 2388 hCoV-19/Denmark/DCGC-9922/2020\|EPI_ISL_669273\|2020 | .................... | .................... | .................... | .................... | .................... | .................... |
| T | 2389 hCoV-19/Denmark/DCGC-9923/2020\|EPI_ISL_669274\|2020 | .................... | .................... | .................... | .................... | .................... | .................... |
| T | 2390 hCoV-19/Denmark/DCGC-9924/2020\|EPI_ISL_669275\|2020 | .................... | .................... | .................... | .................... | .................... | .................... |
| T | 2391 hCoV-19/Denmark/DCGC-9925/2020\|EPI_ISL_669276\|2020 | .................... | .................... | .................... | .................... | .................... | .................... |
| T | 2392 hCoV-19/Denmark/DCGC-9926/2020\|EPI_ISL_669277\|2020 | .................... | .A.................. | .................... | .................... | .................... | ..A................. |
| T | 2393 hCoV-19/Denmark/DCGC-9930/2020\|EPI_ISL_669278\|2020 | .................... | .................... | .................... | .................... | .................... | .................... |
| T | 2394 hCoV-19/Denmark/DCGC-9932/2020\|EPI_ISL_669279\|2020 | .................... | .A.................. | .................... | .................... | .................... | .................... |
| T | 2395 hCoV-19/Denmark/DCGC-9937/2020\|EPI_ISL_669281\|2020 | .................... | .................... | .................... | .................... | .................... | .................... |
| T | 2396 hCoV-19/Denmark/DCGC-9938/2020\|EPI_ISL_669282\|2020 | .................... | .................... | .................... | .................... | .................... | .................... |
| T | 2397 hCoV-19/Denmark/DCGC-9939/2020\|EPI_ISL_669283\|2020 | .................... | .................... | .................... | .................... | .................... | .................... |
| T | 2398 hCoV-19/Denmark/DCGC-9942/2020\|EPI_ISL_669284\|2020 | .................... | .................... | .................... | .................... | .................... | .................... |
| T | 2399 hCoV-19/Denmark/DCGC-9944/2020\|EPI_ISL_669285\|2020 | .................... | .................... | .................... | .................... | .................... | .................... |
| T | 2400 hCoV-19/Denmark/DCGC-9945/2020\|EPI_ISL_669286\|2020 | .................... | .................... | .................... | .................... | .................... | .................... |
| T | 2401 hCoV-19/Denmark/DCGC-9946/2020\|EPI_ISL_669287\|2020 | .................... | .................... | .................... | .................... | .................... | .................... |
| T | 2402 hCoV-19/Denmark/DCGC-9947/2020\|EPI_ISL_669288\|2020 | .................... | ...A................ | .................... | .................... | .................... | .................... |
| T | 2403 hCoV-19/Denmark/DCGC-9949/2020\|EPI_ISL_669289\|2020 | .................... | .................... | .................... | .................... | .................... | .................... |
| T | 2404 hCoV-19/Denmark/DCGC-10774/2020\|EPI_ISL_670200\|202 | .................... | .................... | .................... | .................... | .................... | .................... |
| T | 2405 hCoV-19/Denmark/DCGC-10779/2020\|EPI_ISL_670201\|202 | .................... | .................... | .................... | .................... | .................... | .................... |
| T | 2406 hCoV-19/Denmark/DCGC-10780/2020\|EPI_ISL_670202\|202 | .................... | .................... | .................... | .................... | .................... | .................... |
| T | 2407 hCoV-19/Denmark/DCGC-10781/2020\|EPI_ISL_670203\|202 | .................... | .................... | .................... | .................... | .................... | .................... |
| T | 2408 hCoV-19/Denmark/DCGC-10783/2020\|EPI_ISL_670204\|202 | .................... | .................... | .................... | .................... | .................... | .................... |
| T | 2409 hCoV-19/Denmark/DCGC-10785/2020\|EPI_ISL_670205\|202 | .................... | .................... | .................... | .................... | .................... | .................... |
| T | 2410 hCoV-19/Denmark/DCGC-10786/2020\|EPI_ISL_670206\|202 | .................... | .................... | .................... | .................... | .................... | .................... |
| T | 2411 hCoV-19/Denmark/DCGC-10788/2020\|EPI_ISL_670207\|202 | .................... | .................... | .................... | .................... | .................... | .................... |
| T | 2412 hCoV-19/Denmark/DCGC-10789/2020\|EPI_ISL_670208\|202 | .................... | .................... | .................... | .................... | .................... | .................... |
| T | 2413 hCoV-19/Denmark/DCGC-10790/2020\|EPI_ISL_670209\|202 | .................... | .A.................. | .................... | .................... | .................... | .................... |
| T | 2414 hCoV-19/Denmark/DCGC-10791/2020\|EPI_ISL_670210\|202 | .................... | .................... | .................... | .................... | .................... | .................... |
| T | 2415 hCoV-19/Denmark/DCGC-10792/2020\|EPI_ISL_670211\|202 | .................... | .................... | .................... | .................... | .................... | .................... |
| T | 2416 hCoV-19/Denmark/DCGC-10793/2020\|EPI_ISL_670212\|202 | .................... | .................... | .................... | .................... | .................... | .................... |
| T | 2417 hCoV-19/Denmark/DCGC-10795/2020\|EPI_ISL_670213\|202 | .................... | .................... | .................... | .................... | .................... | .................... |
| T | 2418 hCoV-19/Denmark/DCGC-10796/2020\|EPI_ISL_670214\|202 | .................... | .................... | .................... | .................... | .................... | .................... |
| T | 2419 hCoV-19/Denmark/DCGC-10797/2020\|EPI_ISL_670215\|202 | .................... | .................... | .................... | .................... | .................... | .................... |
| T | 2420 hCoV-19/Denmark/DCGC-10799/2020\|EPI_ISL_670216\|202 | .................... | .................... | .................... | .................... | .................... | .................... |
| T | 2421 hCoV-19/Denmark/DCGC-10801/2020\|EPI_ISL_670217\|202 | .................... | .................... | .................... | .................... | .................... | .................... |
| T | 2422 hCoV-19/Denmark/DCGC-10803/2020\|EPI_ISL_670218\|202 | .................... | .................... | .................... | .................... | .................... | .................... |
| T | 2423 hCoV-19/Denmark/DCGC-10804/2020\|EPI_ISL_670219\|202 | .................... | .................... | .................... | .................... | .................... | .................... |
| T | 2424 hCoV-19/Denmark/DCGC-10806/2020\|EPI_ISL_670220\|202 | .................... | .................... | .................... | .................... | .................... | .................... |
| T | 2425 hCoV-19/Denmark/DCGC-10807/2020\|EPI_ISL_670221\|202 | .................... | .................... | .................... | .................... | .................... | .................... |
| T | 2426 hCoV-19/Denmark/DCGC-10812/2020\|EPI_ISL_670222\|202 | .................... | .................... | .................... | .................... | .................... | .................... |
| T | 2427 hCoV-19/Denmark/DCGC-10813/2020\|EPI_ISL_670223\|202 | .................... | .................... | .................... | .................... | .................... | .................... |
| T | 2428 hCoV-19/Denmark/DCGC-10814/2020\|EPI_ISL_670224\|202 | .................... | .A.................. | .................... | .................... | .................... | .................... |
| T | 2429 hCoV-19/Denmark/DCGC-10815/2020\|EPI_ISL_670225\|202 | .................... | .................... | .................... | .................... | .................... | .................... |
| T | 2430 hCoV-19/Denmark/DCGC-10816/2020\|EPI_ISL_670226\|202 | .................... | .................... | .................... | .................... | .................... | .................... |
| T | 2431 hCoV-19/Denmark/DCGC-10817/2020\|EPI_ISL_670227\|202 | .................... | .................... | .................... | .................... | .................... | .................... |
| T | 2432 hCoV-19/Denmark/DCGC-10820/2020\|EPI_ISL_670228\|202 | .................... | .................... | .................... | .................... | .................... | .................... |
| T | 2433 hCoV-19/Denmark/DCGC-10821/2020\|EPI_ISL_670229\|202 | .................... | .................... | .................... | .................... | .................... | .................... |
| T | 2434 hCoV-19/Denmark/DCGC-10822/2020\|EPI_ISL_670230\|202 | .................... | .................... | .................... | .................... | .................... | .................... |
| T | 2435 hCoV-19/Denmark/DCGC-10823/2020\|EPI_ISL_670231\|202 | .................... | .................... | .................... | .................... | .................... | .................... |
| T | 2436 hCoV-19/Denmark/DCGC-10824/2020\|EPI_ISL_670232\|202 | .................... | .................... | .................... | .................... | .................... | .................... |
| T | 2437 hCoV-19/Denmark/DCGC-10825/2020\|EPI_ISL_670233\|202 | .................... | .................... | .................... | .................... | .................... | .................... |
| T | 2438 hCoV-19/Denmark/DCGC-10827/2020\|EPI_ISL_670234\|202 | .................... | .................... | .................... | .................... | .................... | .................... |
| T | 2439 hCoV-19/Denmark/DCGC-10828/2020\|EPI_ISL_670235\|202 | .................... | .................... | .................... | .................... | .................... | .................... |
| T | 2440 hCoV-19/Denmark/DCGC-10829/2020\|EPI_ISL_670236\|202 | .................... | .................... | .................... | .................... | .................... | .................... |
| T | 2441 hCoV-19/Denmark/DCGC-10830/2020\|EPI_ISL_670237\|202 | .................... | .A.................. | .................... | .................... | .................... | .................... |
| T | 2442 hCoV-19/Denmark/DCGC-10832/2020\|EPI_ISL_670238\|202 | .................... | .................... | .................... | .................... | .................... | .................... |
| T | 2443 hCoV-19/Denmark/DCGC-10833/2020\|EPI_ISL_670239\|202 | .................... | .................... | .................... | .................... | .................... | .................... |

| | | | | | | | | |
|---|---|---|---|---|---|---|---|---|
| T | 2444 hCoV-19/Denmark/DCGC-10834/2020\|EPI_ISL_670240\|202 | ···················· | ···················· | ···················· | ···················· | ···················· | ···················· | |
| T | 2445 hCoV-19/Denmark/DCGC-10835/2020\|EPI_ISL_670241\|202 | ···················· | ···················· | ···················· | ···················· | ···················· | ···················· | |
| T | 2446 hCoV-19/Denmark/DCGC-10836/2020\|EPI_ISL_670242\|202 | ···················· | ···················· | ···················· | ···················· | ···················· | ···················· | |
| T | 2447 hCoV-19/Denmark/DCGC-10837/2020\|EPI_ISL_670243\|202 | ···················· | ···················· | ···················· | ···················· | ···················· | ···················· | |
| T | 2448 hCoV-19/Denmark/DCGC-10838/2020\|EPI_ISL_670244\|202 | ···················· | ···················· | ···················· | ···················· | ···················· | ···················· | |
| T | 2449 hCoV-19/Denmark/DCGC-10839/2020\|EPI_ISL_670245\|202 | ···················· | ···················· | ···················· | ···················· | ···················· | ···················· | |
| T | 2450 hCoV-19/Denmark/DCGC-10840/2020\|EPI_ISL_670246\|202 | ···················· | ···················· | ···················· | ···················· | ···················· | ···················· | |
| T | 2451 hCoV-19/Denmark/DCGC-10841/2020\|EPI_ISL_670247\|202 | ···················· | ···················· | ···················· | ···················· | ···················· | ···················· | |
| T | 2452 hCoV-19/Denmark/DCGC-10845/2020\|EPI_ISL_670248\|202 | ···················· | ···················· | ···················· | ···················· | ···················· | ···················· | |
| T | 2453 hCoV-19/Denmark/DCGC-10847/2020\|EPI_ISL_670249\|202 | ···················· | ···················· | ···················· | ···················· | ···················· | ···················· | |
| T | 2454 hCoV-19/Denmark/DCGC-10848/2020\|EPI_ISL_670250\|202 | ···················· | ···················· | ···················· | ···················· | ···················· | ···················· | |
| T | 2455 hCoV-19/Denmark/DCGC-10853/2020\|EPI_ISL_670251\|202 | ···················· | ···················· | ···················· | ···················· | ···················· | ···················· | |
| T | 2456 hCoV-19/Denmark/DCGC-10857/2020\|EPI_ISL_670252\|202 | ···················· | ···················· | ···················· | ···················· | ···················· | ···················· | |
| T | 2457 hCoV-19/Denmark/DCGC-10858/2020\|EPI_ISL_670253\|202 | ···················· | ···················· | ···················· | ···················· | ···················· | ···················· | |
| T | 2458 hCoV-19/Denmark/DCGC-10866/2020\|EPI_ISL_670254\|202 | ···················· | ···················· | ···················· | ···················· | ···················· | ···················· | |
| T | 2459 hCoV-19/Denmark/DCGC-10867/2020\|EPI_ISL_670255\|202 | ···················· | ·A·················· | ···················· | ···················· | ···················· | ···················· | |
| T | 2460 hCoV-19/Denmark/DCGC-10869/2020\|EPI_ISL_670256\|202 | ···················· | ···················· | ···················· | ···················· | ···················· | ···················· | |
| T | 2461 hCoV-19/Denmark/DCGC-10871/2020\|EPI_ISL_670257\|202 | ···················· | ···················· | ···················· | ···················· | ···················· | ···················· | |
| T | 2462 hCoV-19/Denmark/DCGC-10872/2020\|EPI_ISL_670258\|202 | ···················· | ···················· | ···················· | ···················· | ···················· | ···················· | |
| T | 2463 hCoV-19/Denmark/DCGC-10873/2020\|EPI_ISL_670259\|202 | ···················· | ···················· | ···················· | ···················· | ···················· | ···················· | |
| T | 2464 hCoV-19/Denmark/DCGC-10875/2020\|EPI_ISL_670260\|202 | ···················· | ···················· | ···················· | ···················· | ···················· | ···················· | |
| T | 2465 hCoV-19/Denmark/DCGC-10877/2020\|EPI_ISL_670261\|202 | ···················· | ···················· | ···················· | ···················· | ···················· | ···················· | |
| T | 2466 hCoV-19/Denmark/DCGC-10881/2020\|EPI_ISL_670262\|202 | ···················· | ···················· | ···················· | ···················· | ···················· | ···················· | |
| T | 2467 hCoV-19/Denmark/DCGC-10882/2020\|EPI_ISL_670263\|202 | ···················· | ···················· | ···················· | ···················· | ···················· | ···················· | |
| T | 2468 hCoV-19/Denmark/DCGC-10884/2020\|EPI_ISL_670264\|202 | ···················· | ···················· | ···················· | ···················· | ···················· | ···················· | |
| T | 2469 hCoV-19/Denmark/DCGC-10887/2020\|EPI_ISL_670265\|202 | ···················· | ···················· | ···················· | ···················· | ···················· | ···················· | |
| T | 2470 hCoV-19/Denmark/DCGC-10890/2020\|EPI_ISL_670267\|202 | ···················· | ···················· | ···················· | ···················· | ···················· | ···················· | |
| T | 2471 hCoV-19/Denmark/DCGC-10892/2020\|EPI_ISL_670268\|202 | ···················· | ···················· | ···················· | ···················· | ···················· | ···················· | |
| T | 2472 hCoV-19/Denmark/DCGC-10894/2020\|EPI_ISL_670270\|202 | ···················· | ·A·················· | ···················· | ···················· | ···················· | ···················· | |
| T | 2473 hCoV-19/Denmark/DCGC-10896/2020\|EPI_ISL_670271\|202 | ···················· | ···················· | ···················· | ···················· | ···················· | ···················· | |
| T | 2474 hCoV-19/Denmark/DCGC-10897/2020\|EPI_ISL_670272\|202 | ···················· | ···················· | ···················· | ···················· | ···················· | ···················· | |
| T | 2475 hCoV-19/Denmark/DCGC-10898/2020\|EPI_ISL_670273\|202 | ···················· | ···················· | ···················· | ···················· | ···················· | ···················· | |
| T | 2476 hCoV-19/Denmark/DCGC-10901/2020\|EPI_ISL_670274\|202 | ···················· | ·A·················· | ···················· | ···················· | ···················· | ···················· | |
| T | 2477 hCoV-19/Denmark/DCGC-10902/2020\|EPI_ISL_670275\|202 | ···················· | ···················· | ···················· | ···················· | ···················· | ···················· | |
| T | 2478 hCoV-19/Denmark/DCGC-10927/2020\|EPI_ISL_670287\|202 | ···················· | ···················· | ···················· | ···················· | ···················· | ···················· | |
| T | 2479 hCoV-19/Denmark/DCGC-10928/2020\|EPI_ISL_670288\|202 | ···················· | ···················· | ···················· | ···················· | ···················· | ···················· | |
| T | 2480 hCoV-19/Denmark/DCGC-10929/2020\|EPI_ISL_670289\|202 | ···················· | ···················· | ···················· | ···················· | ···················· | ···················· | |
| T | 2481 hCoV-19/Denmark/DCGC-10939/2020\|EPI_ISL_670298\|202 | ···················· | ···················· | ···················· | ···················· | ···················· | ···················· | |
| T | 2482 hCoV-19/Denmark/DCGC-10940/2020\|EPI_ISL_670299\|202 | ···················· | ···················· | ···················· | ···················· | ···················· | ···················· | |
| T | 2483 hCoV-19/Canada/ON-PHL-20-02208/2020\|EPI_ISL_671520 | ···················· | ···················· | ···················· | ···················· | ···················· | ···················· | |
| T | 2484 hCoV-19/Canada/ON-PHL-20-02291/2020\|EPI_ISL_671521 | ···················· | ···················· | ···················· | ···················· | ···················· | ···················· | |
| T | 2485 hCoV-19/Canada/ON-PHL-20-02292/2020\|EPI_ISL_671522 | ···················· | ···················· | ···················· | ···················· | ···················· | ···················· | |
| T | 2486 hCoV-19/Canada/ON-PHL-20-02295/2020\|EPI_ISL_671523 | ···················· | ···················· | ···················· | ···················· | NNN················· | ···················· | 1 |
| T | 2487 hCoV-19/Canada/ON-PHL-20-02298/2020\|EPI_ISL_671524 | ···················· | ···················· | ···················· | ···················· | ···················· | ···················· | |
| T | 2488 hCoV-19/Canada/ON-PHL-20-02414/2020\|EPI_ISL_671525 | ···················· | ···················· | ···················· | ···················· | ···················· | ···················· | |
| T | 2489 hCoV-19/Canada/ON-PHL-20-02397/2020\|EPI_ISL_671526 | ···················· | ···················· | ···················· | ···················· | ···················· | ···················· | |
| T | 2490 hCoV-19/Canada/ON-PHL-20-02398/2020\|EPI_ISL_671527 | ···················· | ···················· | ···················· | ···················· | ···················· | ···················· | |
| T | 2491 hCoV-19/Canada/ON-PHL-20-02418/2020\|EPI_ISL_671528 | ···················· | ···················· | ···················· | ···················· | ···················· | ···················· | |
| T | 2492 hCoV-19/Canada/ON-PHL-20-02417/2020\|EPI_ISL_671529 | ···················· | ···················· | ···················· | ···················· | ···················· | ···················· | |
| T | 2493 hCoV-19/Canada/ON-PHL-20-02381/2020\|EPI_ISL_671530 | ···················· | ···················· | ···················· | ···················· | ···················· | ···················· | |
| T | 2496 hCoV-19/Canada/ON-PHL-20-02400/2020\|EPI_ISL_671533 | ···················· | ···················· | ···················· | ···················· | ···················· | ···················· | |
| T | 2497 hCoV-19/Canada/ON-PHL-20-02432/2020\|EPI_ISL_671534 | ···················· | ···················· | ···················· | ···················· | ···················· | ···················· | |
| T | 2498 hCoV-19/Canada/ON-PHL-20-02401/2020\|EPI_ISL_671535 | ···················· | ···················· | ···················· | ···················· | ···················· | ···················· | |
| T | 2506 hCoV-19/Canada/ON-PHL-20-02446/2020\|EPI_ISL_671543 | ···················· | ···················· | ··········T········· | ···················· | NNN················· | ···················· | 1 |
| T | 2507 hCoV-19/Canada/ON-PHL-20-02445/2020\|EPI_ISL_671544 | ···················· | ···················· | ···················· | ···················· | ···················· | ···················· | |
| T | 2508 hCoV-19/Canada/ON-PHL-20-02454/2020\|EPI_ISL_671545 | ···················· | ···················· | ···················· | ···················· | ···················· | ···················· | |
| T | 2509 hCoV-19/Canada/ON-PHL-20-02299/2020\|EPI_ISL_671546 | ···················· | ···················· | ···················· | ···················· | ···················· | ···················· | |
| T | 2510 hCoV-19/Canada/ON-PHL-20-02273/2020\|EPI_ISL_671547 | ···················· | ···················· | ···················· | ···················· | ···················· | ···················· | |
| T | 2511 hCoV-19/Canada/ON-PHL-20-02297/2020\|EPI_ISL_671548 | ···················· | ···················· | ···················· | ···················· | ···················· | ···················· | |
| T | 2512 hCoV-19/Canada/ON-PHL-20-02304/2020\|EPI_ISL_671549 | ···················· | ···················· | ···················· | ···················· | ···················· | ···················· | |
| T | 2514 hCoV-19/Canada/ON-PHL-20-02313/2020\|EPI_ISL_671551 | ···················· | ···················· | ···················· | ···················· | ···················· | ···················· | |
| T | 2515 hCoV-19/Canada/ON-PHL-20-02317/2020\|EPI_ISL_671552 | ···················· | ···················· | ···················· | ···················· | ···················· | ···················· | |
| T | 2516 hCoV-19/Canada/ON-PHL-20-02318/2020\|EPI_ISL_671553 | ···················· | ···················· | ···················· | ···················· | ···················· | ···················· | |
| T | 2517 hCoV-19/Canada/ON-PHL-20-02328/2020\|EPI_ISL_671554 | ···················· | ···················· | ···················· | ···················· | ···················· | ···················· | |
| T | 2518 hCoV-19/Canada/ON-PHL-20-02329/2020\|EPI_ISL_671555 | ···················· | ····A··············· | ···················· | ···················· | NNN················· | ···················· | 1 |
| T | 2519 hCoV-19/Canada/ON-PHL-20-02332/2020\|EPI_ISL_671556 | ···················· | ···················· | ···················· | ···················· | ···················· | ···················· | |
| T | 2520 hCoV-19/Canada/ON-PHL-20-02300/2020\|EPI_ISL_671557 | ···················· | ···················· | ···················· | ···················· | ···················· | ···················· | |
| T | 2521 hCoV-19/Canada/ON-PHL-20-02301/2020\|EPI_ISL_671558 | ···················· | ···················· | ···················· | ···················· | ···················· | ···················· | |
| T | 2524 hCoV-19/Canada/ON-PHL-20-02290/2020\|EPI_ISL_671561 | ···················· | ···················· | ···················· | ···················· | ···················· | ···················· | |
| T | 2525 hCoV-19/Canada/ON-PHL-20-02267/2020\|EPI_ISL_671562 | ···················· | ···················· | ···················· | ···················· | ···················· | ···················· | |
| T | 2526 hCoV-19/Canada/ON-PHL-20-02268/2020\|EPI_ISL_671563 | ···················· | ···················· | ···················· | ···················· | ···················· | ···················· | |
| T | 2527 hCoV-19/Canada/ON-PHL-20-02282/2020\|EPI_ISL_671564 | ···················· | ···················· | ···················· | ···················· | ···················· | ···················· | |
| T | 2528 hCoV-19/Canada/ON-PHL-20-02302/2020\|EPI_ISL_671565 | ···················· | ···················· | ···················· | ···················· | ···················· | ···················· | |
| T | 2529 hCoV-19/Canada/ON-PHL-20-02334/2020\|EPI_ISL_671566 | ···················· | ···················· | ···················· | ···················· | ···················· | ···················· | |
| T | 2531 hCoV-19/Canada/ON-PHL-20-02485/2020\|EPI_ISL_671568 | ···················· | ···················· | ···················· | ···················· | N··················· | ···················· | |
| T | 2532 hCoV-19/Canada/ON-PHL-20-02483/2020\|EPI_ISL_671569 | ···················· | ···················· | ···················· | ···················· | ···················· | ···················· | |
| T | 2533 hCoV-19/Canada/ON-PHL-20-02455/2020\|EPI_ISL_671570 | ···················· | ···················· | ···················· | ···················· | ···················· | ···················· | |
| T | 2534 hCoV-19/Canada/ON-PHL-20-02463/2020\|EPI_ISL_671571 | ···················· | ···················· | ···················· | ···················· | ···················· | ···················· | |
| T | 2535 hCoV-19/Canada/ON-PHL-20-02470/2020\|EPI_ISL_671572 | ···················· | ···················· | ···················· | ···················· | ···················· | ···················· | |
| T | 2536 hCoV-19/Canada/ON-PHL-20-02469/2020\|EPI_ISL_671573 | ···················· | ···················· | ···················· | ···················· | ···················· | ···················· | |

| | | | | | | | |
|---|---|---|---|---|---|---|---|
| T | 2537 hCoV-19/Canada/ON-PHL-20-02468/2020\|EPI_ISL_671574 | ···················· | ···················· | ···················· | ···················· | ···················· | ···················· |
| T | 2538 hCoV-19/Canada/ON-PHL-20-02467/2020\|EPI_ISL_671575 | ···················· | ···················· | ···················· | ···················· | ···················· | ···················· |
| T | 2539 hCoV-19/Canada/ON-PHL-20-02466/2020\|EPI_ISL_671576 | ···················· | ···················· | ···················· | ···················· | ···················· | ···················· |
| T | 2542 hCoV-19/Canada/ON-PHL-20-02459/2020\|EPI_ISL_671579 | ···················· | ···················· | ···················· | ···················· | ···················· | ···················· |
| T | 2543 hCoV-19/Canada/ON-PHL-20-02436/2020\|EPI_ISL_671580 | ···················· | ···················· | ···················· | ···················· | ···················· | ···················· |
| T | 2544 hCoV-19/Canada/ON-PHL-20-02437/2020\|EPI_ISL_671581 | ···················· | ···················· | ···················· | ···················· | ···················· | ···················· |
| T | 2546 hCoV-19/Canada/ON-PHL-20-02480/2020\|EPI_ISL_671583 | ···················· | ···················· | ···················· | ···················· | ···················· | ···················· |
| T | 2547 hCoV-19/Canada/ON-PHL-20-02481/2020\|EPI_ISL_671584 | ···················· | ···················· | ···················· | ···················· | ···················· | ···················· |
| T | 2548 hCoV-19/Canada/ON-PHL-20-02479/2020\|EPI_ISL_671585 | ···················· | ···················· | ···················· | ···················· | ···················· | ···················· |
| T | 2551 hCoV-19/Canada/ON-PHL-20-02474/2020\|EPI_ISL_671588 | ···················· | ···················· | ···················· | ···················· | ···················· | ···················· |
| T | 2552 hCoV-19/Canada/ON-PHL-20-02493/2020\|EPI_ISL_671589 | ···················· | ···················· | ···················· | ···················· | ···················· | ···················· |
| T | 2556 hCoV-19/Canada/ON-PHL-20-02430/2020\|EPI_ISL_671593 | ···················· | ···················· | ···················· | ···················· | ···················· | ···················· |
| T | 2560 hCoV-19/Canada/ON-PHL-20-02402/2020\|EPI_ISL_671597 | ···················· | ···················· | ···················· | ···················· | ···················· | ···················· |
| T | 2561 hCoV-19/England/QEUH-AF513E/2020\|EPI_ISL_663001\|20 | ···················· | ·A·················· | ···················· | ···················· | ···················· | ··A················· |
| T | 2562 hCoV-19/England/QEUH-AF48FE/2020\|EPI_ISL_663002\|20 | ···················· | ···················· | ···················· | ···················· | ···················· | ···················· |
| T | 2563 hCoV-19/England/QEUH-AF5022/2020\|EPI_ISL_663004\|20 | ···················· | ···················· | ···················· | ···················· | ···················· | ···················· |
| T | 2564 hCoV-19/England/QEUH-AF570F/2020\|EPI_ISL_663005\|20 | ···················· | ·A·················· | ···················· | ···················· | ···················· | ···················· |
| T | 2565 hCoV-19/England/QEUH-AF4DC2/2020\|EPI_ISL_663006\|20 | ···················· | ···················· | ···················· | ···················· | ···················· | ···················· |
| T | 2566 hCoV-19/England/QEUH-AF53FC/2020\|EPI_ISL_663007\|20 | ···················· | ···················· | ···················· | ···················· | ···················· | ···················· |
| T | 2567 hCoV-19/England/QEUH-AF66F2/2020\|EPI_ISL_663008\|20 | ···················· | ···················· | ···················· | ···················· | ···················· | ···················· |
| T | 2568 hCoV-19/England/QEUH-AF3E84/2020\|EPI_ISL_663009\|20 | ···················· | ···················· | ···················· | ···················· | ···················· | ···················· |
| T | 2569 hCoV-19/England/QEUH-AF6980/2020\|EPI_ISL_663010\|20 | ···················· | ···················· | ···················· | ···················· | ···················· | ···················· |
| T | 2570 hCoV-19/England/QEUH-AF5B6D/2020\|EPI_ISL_663011\|20 | ···················· | ···················· | ···················· | ···················· | ···················· | ···················· |
| T | 2571 hCoV-19/England/QEUH-AF50F5/2020\|EPI_ISL_663012\|20 | ···················· | ···················· | ···················· | ···················· | ···················· | ···················· |
| T | 2572 hCoV-19/England/QEUH-AF5EA0/2020\|EPI_ISL_663013\|20 | ···················· | ···················· | ···················· | ···················· | ···················· | ···················· |
| T | 2573 hCoV-19/England/QEUH-AF4BE6/2020\|EPI_ISL_663014\|20 | ···················· | ···················· | ···················· | ···················· | ···················· | ···················· |
| T | 2574 hCoV-19/England/QEUH-AF4120/2020\|EPI_ISL_663015\|20 | ···················· | ···················· | ···················· | ···················· | ···················· | ···················· |
| T | 2575 hCoV-19/England/QEUH-AF558D/2020\|EPI_ISL_663016\|20 | ···················· | ·A·················· | ···················· | ···················· | ···················· | ··A················· |
| T | 2576 hCoV-19/England/QEUH-AF5E28/2020\|EPI_ISL_663018\|20 | ···················· | ···················· | ···················· | ···················· | ···················· | ···················· |
| T | 2577 hCoV-19/England/QEUH-AF40D8/2020\|EPI_ISL_663019\|20 | ···················· | ···················· | ···················· | ···················· | ···················· | ···················· |
| T | 2578 hCoV-19/England/QEUH-AF4D4A/2020\|EPI_ISL_663020\|20 | ···················· | ···················· | ···················· | ···················· | ·····T·············· | ···················· |
| T | 2579 hCoV-19/England/QEUH-AF50E6/2020\|EPI_ISL_663021\|20 | ···················· | ···················· | ···················· | ···················· | ···················· | ···················· |
| T | 2580 hCoV-19/England/QEUH-AF567B/2020\|EPI_ISL_663022\|20 | ···················· | ···················· | ···················· | ···················· | ···················· | ···················· |
| T | 2581 hCoV-19/England/QEUH-AF4CC5/2020\|EPI_ISL_663023\|20 | ···················· | ···················· | ···················· | ···················· | ···················· | ···················· |
| T | 2582 hCoV-19/England/QEUH-AF40C9/2020\|EPI_ISL_663024\|20 | ···················· | ···················· | ···················· | ···················· | ···················· | ···················· |
| T | 2583 hCoV-19/England/QEUH-AF4919/2020\|EPI_ISL_663025\|20 | ···················· | ·A·················· | ···················· | ···················· | ···················· | ···················· |
| T | 2584 hCoV-19/England/QEUH-AF414E/2020\|EPI_ISL_663026\|20 | ···················· | ···················· | ···················· | ···················· | ···················· | ···················· |
| T | 2585 hCoV-19/England/QEUH-AF69BD/2020\|EPI_ISL_663027\|20 | ···················· | ···················· | ···················· | ···················· | ···················· | ···················· |
| T | 2586 hCoV-19/England/QEUH-AF5778/2020\|EPI_ISL_663029\|20 | ···················· | ···················· | ···················· | ···················· | ···················· | ···················· |
| T | 2587 hCoV-19/England/QEUH-AF4C01/2020\|EPI_ISL_663031\|20 | ···················· | ···················· | ···················· | ···················· | ···················· | ···················· |
| T | 2588 hCoV-19/England/QEUH-AF65F5/2020\|EPI_ISL_663032\|20 | ···················· | ···················· | ···················· | ···················· | ···················· | ···················· |
| T | 2589 hCoV-19/England/QEUH-AF69DB/2020\|EPI_ISL_663033\|20 | ···················· | ···················· | ···················· | ···················· | ···················· | ···················· |
| T | 2590 hCoV-19/England/QEUH-AF52B3/2020\|EPI_ISL_663034\|20 | ···················· | ···················· | ···················· | ···················· | ···················· | ···················· |
| T | 2591 hCoV-19/England/QEUH-AF54CC/2020\|EPI_ISL_663035\|20 | ···················· | ···················· | ···················· | ···················· | ···················· | ···················· |
| T | 2592 hCoV-19/England/QEUH-AF5BA9/2020\|EPI_ISL_663036\|20 | ···················· | ···················· | ···················· | ···················· | ···················· | ···················· |
| T | 2593 hCoV-19/England/QEUH-AF549F/2020\|EPI_ISL_663037\|20 | ···················· | ···················· | ···················· | ···················· | ···················· | ···················· |
| T | 2594 hCoV-19/England/QEUH-AF5E82/2020\|EPI_ISL_663038\|20 | ···················· | ···················· | ···················· | ···················· | ···················· | ···················· |
| T | 2595 hCoV-19/England/QEUH-AF5AD9/2020\|EPI_ISL_663039\|20 | ···················· | ···················· | ···················· | ···················· | ···················· | ···················· |
| T | 2596 hCoV-19/England/QEUH-AF61B5/2020\|EPI_ISL_663041\|20 | ···················· | ···················· | ···················· | ···················· | ···················· | ···················· |
| T | 2597 hCoV-19/England/QEUH-AF4C98/2020\|EPI_ISL_663042\|20 | ···················· | ···················· | ···················· | ···················· | ···················· | ···················· |
| T | 2598 hCoV-19/England/QEUH-AF531A/2020\|EPI_ISL_663043\|20 | ···················· | ···················· | ···················· | ···················· | ···················· | ···················· |
| T | 2599 hCoV-19/England/QEUH-AF5918/2020\|EPI_ISL_663044\|20 | ···················· | ···················· | ···················· | ···················· | ···················· | ···················· |
| T | 2600 hCoV-19/England/QEUH-AF5DEF/2020\|EPI_ISL_663045\|20 | ···················· | ···················· | ···················· | ···················· | ···················· | ···················· |
| T | 2601 hCoV-19/England/QEUH-AF5268/2020\|EPI_ISL_663047\|20 | ···················· | ···················· | ···················· | ···················· | ···················· | ···················· |
| T | 2602 hCoV-19/England/QEUH-AF5E19/2020\|EPI_ISL_663048\|20 | ···················· | ···················· | ···················· | ···················· | ···················· | ···················· |
| T | 2603 hCoV-19/England/QEUH-AF4928/2020\|EPI_ISL_663049\|20 | ···················· | ···················· | ···················· | ···················· | ···················· | ···················· |
| T | 2604 hCoV-19/England/QEUH-AF50C8/2020\|EPI_ISL_663050\|20 | ···················· | ···················· | ···················· | ···················· | ···················· | ···················· |
| T | 2605 hCoV-19/England/QEUH-AF523B/2020\|EPI_ISL_663052\|20 | ···················· | ···················· | ···················· | ···················· | ···················· | ···················· |
| T | 2606 hCoV-19/England/QEUH-AF3DD2/2020\|EPI_ISL_663053\|20 | ···················· | ···················· | ···················· | ···················· | ···················· | ···················· |
| T | 2607 hCoV-19/England/QEUH-AF407E/2020\|EPI_ISL_663054\|20 | ···················· | ···················· | ···················· | ···················· | ···················· | ···················· |
| T | 2608 hCoV-19/England/QEUH-AF580C/2020\|EPI_ISL_663056\|20 | ···················· | ···················· | ···················· | ···················· | ···················· | ···················· |
| T | 2609 hCoV-19/England/QEUH-AF5AE8/2020\|EPI_ISL_663057\|20 | ···················· | ···················· | ···················· | ···················· | ···················· | ···················· |
| T | 2610 hCoV-19/England/QEUH-AF58B1/2020\|EPI_ISL_663058\|20 | ···················· | ···················· | ···················· | ···················· | ···················· | ···················· |
| T | 2611 hCoV-19/England/QEUH-AF5E64/2020\|EPI_ISL_663060\|20 | ···················· | ···················· | ···················· | ···················· | ···················· | ···················· |
| T | 2612 hCoV-19/England/QEUH-AF57D2/2020\|EPI_ISL_663063\|20 | ···················· | ·A·················· | ···················· | ···················· | ···················· | ···················· |
| T | 2613 hCoV-19/England/QEUH-AF408D/2020\|EPI_ISL_663064\|20 | ···················· | ···················· | ···················· | ···················· | ···················· | ···················· |
| T | 2614 hCoV-19/England/QEUH-AF622B/2020\|EPI_ISL_663065\|20 | ···················· | ···················· | ···················· | ···················· | ···················· | ···················· |
| T | 2615 hCoV-19/England/QEUH-AF505F/2020\|EPI_ISL_663066\|20 | ···················· | ···················· | ···················· | ···················· | ···················· | ···················· |
| T | 2616 hCoV-19/England/QEUH-AF699F/2020\|EPI_ISL_663068\|20 | ···················· | ···················· | ···················· | ···················· | ···················· | ···················· |
| T | 2617 hCoV-19/England/QEUH-AF3DB4/2020\|EPI_ISL_663069\|20 | ···················· | ···················· | ···················· | ···················· | ···················· | ···················· |
| T | 2618 hCoV-19/England/QEUH-AF5BC7/2020\|EPI_ISL_663070\|20 | ···················· | ···················· | ···················· | ···················· | ···················· | ···················· |
| T | 2619 hCoV-19/England/QEUH-AF3D69/2020\|EPI_ISL_663071\|20 | ···················· | ···················· | ···················· | ···················· | ···················· | ···················· |
| T | 2620 hCoV-19/England/QEUH-AF6249/2020\|EPI_ISL_663072\|20 | ···················· | ···················· | ···················· | ···················· | ···················· | ···················· |
| T | 2621 hCoV-19/England/QEUH-AF616A/2020\|EPI_ISL_663074\|20 | ···················· | ···················· | ···················· | ···················· | ···················· | ···················· |
| T | 2622 hCoV-19/England/QEUH-AF50D7/2020\|EPI_ISL_663075\|20 | ···················· | ···················· | ···················· | ···················· | ···················· | ···················· |
| T | 2623 hCoV-19/England/QEUH-AF58DF/2020\|EPI_ISL_663078\|20 | ···················· | ···················· | ···················· | ···················· | ···················· | ···················· |
| T | 2624 hCoV-19/England/QEUH-AF5329/2020\|EPI_ISL_663079\|20 | ···················· | ···················· | ···················· | ···················· | ···················· | ···················· |
| T | 2625 hCoV-19/England/QEUH-AF3D5A/2020\|EPI_ISL_663080\|20 | ···················· | ···················· | ···················· | ···················· | ···················· | ···················· |
| T | 2626 hCoV-19/England/QEUH-AF55E7/2020\|EPI_ISL_663081\|20 | ···················· | ···················· | ···················· | ···················· | ···················· | ···················· |
| T | 2627 hCoV-19/England/QEUH-AF5B4F/2020\|EPI_ISL_663082\|20 | ···················· | ···················· | ···················· | ···················· | ···················· | ···················· |

| | | | | | | | |
|---|---|---|---|---|---|---|---|
| T | 2628 hCoV-19/England/QEUH-AF574B/2020\|EPI_ISL_663083\|20 | ···················· | ···················· | ···················· | ···················· | ···················· | ···················· |
| T | 2629 hCoV-19/England/QEUH-AF66B6/2020\|EPI_ISL_663084\|20 | ···················· | ···················· | ···················· | ···················· | ···················· | ···················· |
| T | 2630 hCoV-19/England/QEUH-AF50B9/2020\|EPI_ISL_663085\|20 | ···················· | ···················· | ···················· | ···················· | ···················· | ···················· |
| T | 2631 hCoV-19/England/QEUH-AF4885/2020\|EPI_ISL_663086\|20 | ···················· | ···················· | ···················· | ···················· | ···················· | ···················· |
| T | 2632 hCoV-19/England/QEUH-AF5B8B/2020\|EPI_ISL_663087\|20 | ···················· | ···················· | ···················· | ···················· | ···················· | ···················· |
| T | 2633 hCoV-19/England/QEUH-AF3DE1/2020\|EPI_ISL_663088\|20 | ···················· | ···················· | ···················· | ···················· | ···················· | ···················· |
| T | 2634 hCoV-19/England/QEUH-AF54F9/2020\|EPI_ISL_663089\|20 | ···················· | ···················· | ···················· | ···················· | ···················· | ···················· |
| T | 2635 hCoV-19/England/QEUH-AF5848/2020\|EPI_ISL_663090\|20 | ···················· | ···················· | ···················· | ···················· | ···················· | ···················· |
| T | 2636 hCoV-19/England/QEUH-AF5893/2020\|EPI_ISL_663091\|20 | ···················· | ···················· | ···················· | ···················· | ···················· | ···················· |
| T | 2637 hCoV-19/England/QEUH-AF5875/2020\|EPI_ISL_663092\|20 | ···················· | ···················· | ···················· | ···················· | ···················· | ···················· |
| T | 2639 hCoV-19/England/QEUH-AF662F/2020\|EPI_ISL_663094\|20 | ···················· | ···················· | ···················· | ···················· | ···················· | ···················· |
| T | 2640 hCoV-19/England/QEUH-AF52A4/2020\|EPI_ISL_663095\|20 | ···················· | ···················· | ···················· | ···················· | ···················· | ···················· |
| T | 2641 hCoV-19/England/QEUH-AF4B6E/2020\|EPI_ISL_663096\|20 | ···················· | ···················· | ···················· | ···················· | ······T············· | ···················· |
| T | 2642 hCoV-19/England/QEUH-AF5435/2020\|EPI_ISL_663097\|20 | ···················· | ···················· | ···················· | ···················· | ···················· | ···················· |
| T | 2643 hCoV-19/England/QEUH-AF5480/2020\|EPI_ISL_663098\|20 | ···················· | ···················· | ···················· | ···················· | ···················· | ···················· |
| T | 2644 hCoV-19/England/QEUH-AF57C3/2020\|EPI_ISL_663099\|20 | ···················· | ···················· | ···················· | ···················· | ···················· | ···················· |
| T | 2645 hCoV-19/Scotland/EDB9387/2020\|EPI_ISL_665646\|2020- | ···················· | ···················· | ···················· | ···················· | ···················· | ···················· |
| T | 2646 hCoV-19/Wales/PHWC-48CBE9/2020\|EPI_ISL_665647\|2020 | ···················· | ···················· | ···················· | ···················· | ···················· | ···················· |
| T | 2647 hCoV-19/England/LOND-12513B7/2020\|EPI_ISL_665648\|2 | ···················· | ···················· | ···················· | ···················· | ···················· | ···················· |
| T | 2648 hCoV-19/England/LOND-1250F4B/2020\|EPI_ISL_665649\|2 | ···················· | ·A·················· | ···················· | ···················· | ···················· | ···A················ |
| T | 2649 hCoV-19/England/NOTT-11D825/2020\|EPI_ISL_665650\|20 | ···················· | ·A·················· | ···················· | ···················· | ···················· | ···A················ |
| T | 2650 hCoV-19/England/QEUH-AF1590/2020\|EPI_ISL_665651\|20 | ···················· | ·A·················· | ···················· | ···················· | ···················· | ···A················ |
| T | 2651 hCoV-19/England/MILK-A23CAF/2020\|EPI_ISL_665652\|20 | ···················· | ···················· | ···················· | ···················· | ···················· | ···················· |
| T | 2652 hCoV-19/Wales/PHWC-48B7E6/2020\|EPI_ISL_665653\|2020 | ···················· | ···················· | ···················· | ···················· | ···················· | ···················· |
| T | 2653 hCoV-19/Wales/PHWC-48CB8F/2020\|EPI_ISL_665654\|2020 | ···················· | ···················· | ···················· | ···················· | ···················· | ···················· |
| T | 2654 hCoV-19/Wales/PHWC-48CC31/2020\|EPI_ISL_665655\|2020 | ···················· | ···················· | ···················· | ···················· | ···················· | ···················· |
| T | 2655 hCoV-19/USA/CA-ALSR-4510/2020\|EPI_ISL_666976\|2020- | ···················· | ···················· | ···················· | ···················· | ···················· | ···············A···· |
| T | 2656 hCoV-19/USA/CA-ALSR-4512/2020\|EPI_ISL_666977\|2020- | ···················· | ·A·················· | ···················· | ···················· | ···················· | ···················· |
| T | 2657 hCoV-19/USA/CA-ALSR-4513/2020\|EPI_ISL_666978\|2020- | ···················· | ·A·················· | ···················· | ···················· | ···················· | ···················· |
| T | 2658 hCoV-19/USA/CA-ALSR-4515/2020\|EPI_ISL_666979\|2020- | ···················· | ···················· | ···················· | ···················· | ···················· | ···················· |
| T | 2659 hCoV-19/USA/CA-ALSR-4516/2020\|EPI_ISL_666980\|2020- | ···················· | ···················· | ···················· | ···················· | ···················· | ···················· |
| T | 2660 hCoV-19/USA/CA-ALSR-4517/2020\|EPI_ISL_666981\|2020- | ···T················ | ·A·················· | ···················· | ···················· | ···················· | ···················· |
| T | 2661 hCoV-19/USA/CA-ALSR-4519/2020\|EPI_ISL_666982\|2020- | ···················· | ···················· | ···················· | ···················· | ···················· | ···················· |
| T | 2662 hCoV-19/USA/CA-ALSR-4523/2020\|EPI_ISL_666984\|2020- | ···················· | ·A·················· | ···················· | ···················· | ···················· | ···················· |
| T | 2663 hCoV-19/USA/CA-ALSR-4524/2020\|EPI_ISL_666985\|2020- | ···················· | ···················· | ···················· | ···················· | ···················· | ···················· |
| T | 2664 hCoV-19/USA/CA-ALSR-4525/2020\|EPI_ISL_666986\|2020- | ···················· | ···················· | ···················· | ···················· | ···················· | ···················· |
| T | 2665 hCoV-19/USA/CA-ALSR-4527/2020\|EPI_ISL_666987\|2020- | ···················· | ···················· | ···················· | ···················· | ···················· | ···················· |
| T | 2666 hCoV-19/USA/CA-ALSR-4529/2020\|EPI_ISL_666989\|2020- | ···················· | ·A·················· | ···················· | ···················· | ···················· | ···················· |
| T | 2667 hCoV-19/USA/CA-ALSR-4530/2020\|EPI_ISL_666990\|2020- | ···················· | ···················· | ···················· | ···················· | ···················· | ···················· |
| T | 2668 hCoV-19/USA/CA-ALSR-4533/2020\|EPI_ISL_666992\|2020- | ···················· | ·A·················· | ···················· | ···················· | ···················· | ···················· |
| T | 2669 hCoV-19/USA/CA-ALSR-4534/2020\|EPI_ISL_666993\|2020- | ···················· | ·A·················· | ···················· | ···················· | ···················· | ···················· |
| T | 2670 hCoV-19/USA/CA-ALSR-4535/2020\|EPI_ISL_666994\|2020- | ···················· | ·A·················· | ···················· | ···················· | ···················· | ···················· |
| T | 2671 hCoV-19/USA/CA-ALSR-4536/2020\|EPI_ISL_666995\|2020- | ···················· | ···················· | ···················· | ···················· | ···················· | ···················· |
| T | 2672 hCoV-19/USA/CA-ALSR-4537/2020\|EPI_ISL_666996\|2020- | ···················· | ·A·················· | ···················· | ···················· | ···················· | ···················· |
| T | 2673 hCoV-19/USA/CA-ALSR-4539/2020\|EPI_ISL_666997\|2020- | ···················· | ···················· | ···················· | ···················· | ···················· | ···················· |
| T | 2674 hCoV-19/USA/CA-ALSR-4540/2020\|EPI_ISL_666998\|2020- | ···················· | ···················· | ···················· | ···················· | ···················· | ···················· |
| T | 2675 hCoV-19/USA/CA-ALSR-4541/2020\|EPI_ISL_666999\|2020- | ···················· | ···················· | ···················· | ···················· | ···················· | ···················· |
| T | 2676 hCoV-19/Denmark/DCGC-9744/2020\|EPI_ISL_669141\|2020 | ···················· | ···················· | ···················· | ···················· | ···················· | ···················· |
| T | 2677 hCoV-19/Denmark/DCGC-9757/2020\|EPI_ISL_669150\|2020 | ···················· | ···················· | ···················· | ···················· | ···················· | ···················· |
| T | 2678 hCoV-19/Denmark/DCGC-9758/2020\|EPI_ISL_669151\|2020 | ···················· | ···················· | ···················· | ···················· | ···················· | ···················· |
| T | 2679 hCoV-19/Denmark/DCGC-9759/2020\|EPI_ISL_669152\|2020 | ···················· | ···················· | ···················· | ···················· | ···················· | ···················· |
| T | 2680 hCoV-19/Denmark/DCGC-9760/2020\|EPI_ISL_669153\|2020 | ···················· | ···················· | ···················· | ···················· | ···················· | ···················· |
| T | 2681 hCoV-19/Denmark/DCGC-9761/2020\|EPI_ISL_669154\|2020 | ···················· | ···················· | ···················· | ···················· | ···················· | ···················· |
| T | 2682 hCoV-19/Denmark/DCGC-9762/2020\|EPI_ISL_669155\|2020 | ···················· | ···················· | ···················· | ···················· | ···················· | ···················· |
| T | 2683 hCoV-19/Denmark/DCGC-9763/2020\|EPI_ISL_669156\|2020 | ···················· | ···················· | ···················· | ···················· | ······T············· | ···················· |
| T | 2684 hCoV-19/Denmark/DCGC-9764/2020\|EPI_ISL_669157\|2020 | ···················· | ···················· | ···················· | ···················· | ···················· | ···················· |
| T | 2685 hCoV-19/Denmark/DCGC-9765/2020\|EPI_ISL_669158\|2020 | ···················· | ···················· | ···················· | ···················· | ···················· | ···················· |
| T | 2686 hCoV-19/Denmark/DCGC-9766/2020\|EPI_ISL_669159\|2020 | ···················· | ···················· | ···················· | ···················· | ···················· | ···················· |
| T | 2687 hCoV-19/Denmark/DCGC-9767/2020\|EPI_ISL_669160\|2020 | ···················· | ···················· | ···················· | ···················· | ···················· | ···················· |
| T | 2688 hCoV-19/Denmark/DCGC-9769/2020\|EPI_ISL_669161\|2020 | ···················· | ···················· | ···················· | ···················· | ···················· | ···················· |
| T | 2689 hCoV-19/Denmark/DCGC-9770/2020\|EPI_ISL_669162\|2020 | ···················· | ···················· | ···················· | ···················· | ···················· | ···················· |
| T | 2690 hCoV-19/Denmark/DCGC-9771/2020\|EPI_ISL_669163\|2020 | ···················· | ···················· | ···················· | ···················· | ···················· | ···················· |
| T | 2691 hCoV-19/Denmark/DCGC-9772/2020\|EPI_ISL_669164\|2020 | ···················· | ···················· | ···················· | ···················· | ···················· | ···················· |
| T | 2692 hCoV-19/Denmark/DCGC-9773/2020\|EPI_ISL_669165\|2020 | ···················· | ···················· | ···················· | ···················· | ···················· | ···················· |
| T | 2693 hCoV-19/Denmark/DCGC-9774/2020\|EPI_ISL_669166\|2020 | ···················· | ···················· | ···················· | ···················· | ···················· | ···················· |
| T | 2694 hCoV-19/Denmark/DCGC-9775/2020\|EPI_ISL_669167\|2020 | ···················· | ···················· | ···················· | ···················· | ···················· | ···················· |
| T | 2695 hCoV-19/Denmark/DCGC-9776/2020\|EPI_ISL_669168\|2020 | ···················· | ···················· | ···················· | ···················· | ···················· | ···················· |
| T | 2696 hCoV-19/Denmark/DCGC-9777/2020\|EPI_ISL_669169\|2020 | ···················· | ···················· | ···················· | ···················· | ···················· | ···················· |
| T | 2697 hCoV-19/Denmark/DCGC-9814/2020\|EPI_ISL_669200\|2020 | ···················· | ···················· | ···················· | ···················· | ···················· | ···················· |
| T | 2698 hCoV-19/Denmark/DCGC-9815/2020\|EPI_ISL_669201\|2020 | ···················· | ···················· | ···················· | ···················· | ···················· | ···················· |
| T | 2699 hCoV-19/Denmark/DCGC-9820/2020\|EPI_ISL_669202\|2020 | ···················· | ···················· | ···················· | ···················· | ···················· | ···················· |
| T | 2700 hCoV-19/Denmark/DCGC-9821/2020\|EPI_ISL_669203\|2020 | ···················· | ···················· | ···················· | ···················· | ···················· | ···················· |
| T | 2701 hCoV-19/Denmark/DCGC-9823/2020\|EPI_ISL_669204\|2020 | ···················· | ···················· | ···················· | ···················· | ···················· | ···················· |
| T | 2702 hCoV-19/Denmark/DCGC-9825/2020\|EPI_ISL_669205\|2020 | ···················· | ···················· | ···················· | ···················· | ···················· | ···················· |
| T | 2703 hCoV-19/Denmark/DCGC-9826/2020\|EPI_ISL_669206\|2020 | ···················· | ···················· | ···················· | ···················· | ···················· | ···················· |
| T | 2704 hCoV-19/Denmark/DCGC-9827/2020\|EPI_ISL_669207\|2020 | ···················· | ···················· | ···················· | ···················· | ···················· | ···················· |
| T | 2705 hCoV-19/Denmark/DCGC-9830/2020\|EPI_ISL_669208\|2020 | ···················· | ···················· | ···················· | ···················· | ···················· | ···················· |
| T | 2706 hCoV-19/Denmark/DCGC-9951/2020\|EPI_ISL_669290\|2020 | ···················· | ···················· | ···················· | ···················· | ···················· | ···················· |
| T | 2707 hCoV-19/Denmark/DCGC-9954/2020\|EPI_ISL_669291\|2020 | ···················· | ···················· | ···················· | ···················· | ···················· | ···················· |
| T | 2708 hCoV-19/Denmark/DCGC-9955/2020\|EPI_ISL_669292\|2020 | ···················· | ···················· | ···················· | ···················· | ···················· | ···················· |

| | | | | | | | |
|---|---|---|---|---|---|---|---|
| T | 2709 hCoV-19/Denmark/DCGC-9956/2020\|EPI_ISL_669293\|2020 | ···················· | ···················· | ···················· | ···················· | ···················· | ···················· |
| T | 2710 hCoV-19/Denmark/DCGC-9958/2020\|EPI_ISL_669294\|2020 | ···················· | ···················· | ···················· | ···················· | ···················· | ···················· |
| T | 2711 hCoV-19/Denmark/DCGC-9959/2020\|EPI_ISL_669295\|2020 | ···················· | ···················· | ···················· | ···················· | ···················· | ···················· |
| T | 2712 hCoV-19/Denmark/DCGC-9960/2020\|EPI_ISL_669296\|2020 | ···················· | ···················· | ···················· | ···················· | ···················· | ···················· |
| T | 2713 hCoV-19/Denmark/DCGC-9963/2020\|EPI_ISL_669297\|2020 | ···················· | ···················· | ···················· | ···················· | ···················· | ···················· |
| T | 2714 hCoV-19/Denmark/DCGC-9964/2020\|EPI_ISL_669298\|2020 | ···················· | ···················· | ···················· | ···················· | ···················· | ···················· |
| T | 2715 hCoV-19/Denmark/DCGC-9965/2020\|EPI_ISL_669299\|2020 | ···················· | ···················· | ···················· | ···················· | ···················· | ···················· |
| T | 2716 hCoV-19/Denmark/DCGC-10663/2020\|EPI_ISL_670122\|202 | ···················· | ···················· | ···················· | ···················· | ···················· | ···················· |
| T | 2717 hCoV-19/Denmark/DCGC-10664/2020\|EPI_ISL_670123\|202 | ···················· | ···················· | ···················· | ···················· | ···················· | ···················· |
| T | 2718 hCoV-19/Denmark/DCGC-10668/2020\|EPI_ISL_670124\|202 | ···················· | ···················· | ···················· | ···················· | ···················· | ···················· |
| T | 2719 hCoV-19/Denmark/DCGC-10671/2020\|EPI_ISL_670125\|202 | ···················· | ···················· | ···················· | ···················· | ···················· | ···················· |
| T | 2720 hCoV-19/Denmark/DCGC-10673/2020\|EPI_ISL_670126\|202 | ···················· | ···················· | ···················· | ···················· | ···················· | ···················· |
| T | 2721 hCoV-19/Denmark/DCGC-10674/2020\|EPI_ISL_670127\|202 | ···················· | ···················· | ···················· | ···················· | ···················· | ···················· |
| T | 2722 hCoV-19/Denmark/DCGC-10675/2020\|EPI_ISL_670128\|202 | ···················· | ···················· | ···················· | ···················· | ···················· | ···················· |
| T | 2723 hCoV-19/Denmark/DCGC-10676/2020\|EPI_ISL_670129\|202 | ···················· | ···················· | ···················· | ···················· | ···················· | ···················· |
| T | 2724 hCoV-19/Denmark/DCGC-10678/2020\|EPI_ISL_670130\|202 | ···················· | ···················· | ···················· | ···················· | ···················· | ···················· |
| T | 2725 hCoV-19/Denmark/DCGC-10681/2020\|EPI_ISL_670133\|202 | ···················· | ···················· | ···················· | ···················· | ···················· | ···················· |
| T | 2726 hCoV-19/Denmark/DCGC-10682/2020\|EPI_ISL_670134\|202 | ···················· | ···················· | ···················· | ···················· | ···················· | ···················· |
| T | 2727 hCoV-19/Denmark/DCGC-10683/2020\|EPI_ISL_670135\|202 | ···················· | ·A·················· | ···················· | ···················· | ···················· | ···················· |
| T | 2728 hCoV-19/Denmark/DCGC-10684/2020\|EPI_ISL_670136\|202 | ···················· | ···················· | ···················· | ···················· | ···················· | ···················· |
| T | 2729 hCoV-19/Denmark/DCGC-10685/2020\|EPI_ISL_670137\|202 | ···················· | ···················· | ···················· | ···················· | ···················· | ···················· |
| T | 2730 hCoV-19/Denmark/DCGC-10687/2020\|EPI_ISL_670138\|202 | ···················· | ···················· | ···················· | ···················· | ···················· | ···················· |
| T | 2731 hCoV-19/Denmark/DCGC-10689/2020\|EPI_ISL_670139\|202 | ···················· | ···················· | ···················· | ···················· | ···················· | ···················· |
| T | 2732 hCoV-19/Denmark/DCGC-10690/2020\|EPI_ISL_670140\|202 | ···················· | ···················· | ···················· | ···················· | ···················· | ···················· |
| T | 2733 hCoV-19/Denmark/DCGC-10691/2020\|EPI_ISL_670141\|202 | ···················· | ···················· | ···················· | ···················· | ···················· | ···················· |
| T | 2734 hCoV-19/Denmark/DCGC-10692/2020\|EPI_ISL_670142\|202 | ···················· | ···················· | ···················· | ···················· | ···················· | ···················· |
| T | 2735 hCoV-19/Denmark/DCGC-10696/2020\|EPI_ISL_670143\|202 | ···················· | ···················· | ···················· | ···················· | ···················· | ···················· |
| T | 2736 hCoV-19/Denmark/DCGC-10698/2020\|EPI_ISL_670144\|202 | ···················· | ···················· | ···················· | ···················· | ···················· | ···················· |
| T | 2737 hCoV-19/Denmark/DCGC-10699/2020\|EPI_ISL_670145\|202 | ···················· | ···················· | ···················· | ···················· | ···················· | ···················· |
| T | 2738 hCoV-19/Denmark/DCGC-10700/2020\|EPI_ISL_670146\|202 | ···················· | ···················· | ···················· | ···················· | ···················· | ···················· |
| T | 2739 hCoV-19/Denmark/DCGC-10701/2020\|EPI_ISL_670147\|202 | ···················· | ···················· | ···················· | ···················· | ···················· | ···················· |
| T | 2740 hCoV-19/Denmark/DCGC-10702/2020\|EPI_ISL_670148\|202 | ···················· | ···················· | ···················· | ···················· | ···················· | ···················· |
| T | 2741 hCoV-19/Denmark/DCGC-10704/2020\|EPI_ISL_670149\|202 | ···················· | ···················· | ···················· | ···················· | ···················· | ···················· |
| T | 2742 hCoV-19/Denmark/DCGC-10705/2020\|EPI_ISL_670150\|202 | ···················· | ···················· | ···················· | ···················· | ······T············· | ···················· |
| T | 2743 hCoV-19/Denmark/DCGC-10707/2020\|EPI_ISL_670151\|202 | ···················· | ···················· | ···················· | ···················· | ···················· | ···················· |
| T | 2744 hCoV-19/Denmark/DCGC-10708/2020\|EPI_ISL_670152\|202 | ···················· | ···················· | ···················· | ···················· | ···················· | ···················· |
| T | 2745 hCoV-19/Denmark/DCGC-10709/2020\|EPI_ISL_670153\|202 | ···················· | ···················· | ···················· | ···················· | ···················· | ···················· |
| T | 2746 hCoV-19/Denmark/DCGC-10710/2020\|EPI_ISL_670154\|202 | ···················· | ···················· | ···················· | ···················· | ···················· | ···················· |
| T | 2747 hCoV-19/Denmark/DCGC-10904/2020\|EPI_ISL_670276\|202 | ···················· | ···················· | ···················· | ···················· | ···················· | ···················· |
| T | 2748 hCoV-19/Denmark/DCGC-10905/2020\|EPI_ISL_670277\|202 | ···················· | ···················· | ···················· | ···················· | ···················· | ···················· |
| T | 2749 hCoV-19/Denmark/DCGC-10906/2020\|EPI_ISL_670278\|202 | ···················· | ···················· | ···················· | ···················· | ···················· | ···················· |
| T | 2750 hCoV-19/Denmark/DCGC-10909/2020\|EPI_ISL_670279\|202 | ···················· | ···················· | ···················· | ···················· | ···················· | ···················· |
| T | 2751 hCoV-19/Denmark/DCGC-10911/2020\|EPI_ISL_670280\|202 | ···················· | ···················· | ···················· | ···················· | ···················· | ···················· |
| T | 2752 hCoV-19/Denmark/DCGC-10913/2020\|EPI_ISL_670281\|202 | ···················· | ···················· | ···················· | ···················· | ···················· | ···················· |
| T | 2753 hCoV-19/Denmark/DCGC-10919/2020\|EPI_ISL_670282\|202 | ···················· | ···················· | ···················· | ···················· | ···················· | ···················· |
| T | 2754 hCoV-19/Denmark/DCGC-10920/2020\|EPI_ISL_670283\|202 | ···················· | ···················· | ···················· | ···················· | ···················· | ···················· |
| T | 2755 hCoV-19/Denmark/DCGC-10921/2020\|EPI_ISL_670284\|202 | ···················· | ···················· | ···················· | ···················· | ···················· | ···················· |
| T | 2756 hCoV-19/Denmark/DCGC-10922/2020\|EPI_ISL_670285\|202 | ···················· | ···················· | ···················· | ···················· | ···················· | ···················· |
| T | 2757 hCoV-19/Denmark/DCGC-10925/2020\|EPI_ISL_670286\|202 | ···················· | ···················· | ···················· | ···················· | ···················· | ···················· |
| T | 2758 hCoV-19/Denmark/DCGC-10930/2020\|EPI_ISL_670290\|202 | ···················· | ···················· | ···················· | ···················· | ···················· | ···················· |
| T | 2759 hCoV-19/Denmark/DCGC-10931/2020\|EPI_ISL_670291\|202 | ···················· | ···················· | ···················· | ···················· | ···················· | ···················· |
| T | 2760 hCoV-19/Denmark/DCGC-10932/2020\|EPI_ISL_670292\|202 | ···················· | ···················· | ···················· | ···················· | ···················· | ···················· |
| T | 2761 hCoV-19/Denmark/DCGC-10933/2020\|EPI_ISL_670293\|202 | ···················· | ·A·················· | ···················· | ···················· | ···················· | ···················· |
| T | 2762 hCoV-19/Denmark/DCGC-10934/2020\|EPI_ISL_670294\|202 | ···················· | ···················· | ···················· | ···················· | ···················· | ···················· |
| T | 2763 hCoV-19/Denmark/DCGC-10935/2020\|EPI_ISL_670295\|202 | ···················· | ···················· | ···················· | ···················· | ···················· | ···················· |
| T | 2764 hCoV-19/Denmark/DCGC-10937/2020\|EPI_ISL_670296\|202 | ···················· | ···················· | ···················· | ···················· | ···················· | ···················· |
| T | 2765 hCoV-19/Denmark/DCGC-10938/2020\|EPI_ISL_670297\|202 | ···················· | ···················· | ···················· | ···················· | ···················· | ···················· |
| T | 2766 hCoV-19/Hungary/UD-58450/2020\|EPI_ISL_671453\|2020- | ···················· | ···················· | ···················· | ···················· | ···················· | ···················· |
| T | 2767 hCoV-19/Hungary/UD-58469/2020\|EPI_ISL_671454\|2020- | ···················· | ···················· | ···················· | ···················· | ···················· | ···················· |
| T | 2768 hCoV-19/Hungary/UD-90259/2020\|EPI_ISL_671455\|2020- | ···················· | ···················· | ···················· | ···················· | ···················· | ···················· |
| T | 2769 hCoV-19/Hungary/UD-90279/2020\|EPI_ISL_671456\|2020- | ···················· | ···················· | ···················· | ···················· | ···················· | ···················· |
| T | 2770 hCoV-19/Hungary/UD-90286/2020\|EPI_ISL_671457\|2020- | ···················· | ···················· | ···················· | ···················· | ···················· | ···················· |
| T | 2771 hCoV-19/Hungary/UD-90300/2020\|EPI_ISL_671458\|2020- | ···················· | ···················· | ···················· | ···················· | ···················· | ···················· |
| T | 2772 hCoV-19/Hungary/UD-90319/2020\|EPI_ISL_671459\|2020- | ···················· | ···················· | ···················· | ···················· | ···················· | ···················· |
| T | 2773 hCoV-19/Hungary/UD-90373/2020\|EPI_ISL_671462\|2020- | ···················· | ···················· | ···················· | ···················· | ···················· | ···················· |
| T | 2774 hCoV-19/Hungary/UD-90381/2020\|EPI_ISL_671463\|2020- | ···················· | ···················· | ···················· | ···················· | ···················· | ···················· |
| T | 2775 hCoV-19/Hungary/UD-90383/2020\|EPI_ISL_671464\|2020- | ···················· | ···················· | ···················· | ···················· | ···················· | ···················· |
| T | 2776 hCoV-19/Hungary/UD-90389/2020\|EPI_ISL_671465\|2020- | ···················· | ···················· | ···················· | ···················· | ···················· | ···················· |
| T | 2777 hCoV-19/Hungary/UD-90395/2020\|EPI_ISL_671466\|2020- | ···················· | ···················· | ···················· | ···················· | ···················· | ···················· |
| T | 2778 hCoV-19/Hungary/UD-90402/2020\|EPI_ISL_671467\|2020- | ···················· | ···················· | ···················· | ···················· | ···················· | ···················· |
| T | 2779 hCoV-19/Hungary/UD-90448/2020\|EPI_ISL_671468\|2020- | ···················· | ···················· | ···················· | ···················· | ···················· | ···················· |
| T | 2780 hCoV-19/Hungary/UD-90455/2020\|EPI_ISL_671469\|2020- | ···················· | ···················· | ···················· | ···················· | ···················· | ···················· |
| T | 2781 hCoV-19/Hungary/UD-90462/2020\|EPI_ISL_671470\|2020- | ···················· | ···················· | ···················· | ···················· | ···················· | ···················· |
| T | 2782 hCoV-19/Hungary/UD-90469/2020\|EPI_ISL_671471\|2020- | ···················· | ···················· | ···················· | ···················· | ···················· | ···················· |
| T | 2783 hCoV-19/Hungary/UD-90491/2020\|EPI_ISL_671472\|2020- | ···················· | ···················· | ···················· | ···················· | ···················· | ···················· |
| T | 2784 hCoV-19/Hungary/UD-90512/2020\|EPI_ISL_671473\|2020- | ···················· | ···················· | ···················· | ···················· | ···················· | ···················· |
| T | 2785 hCoV-19/Hungary/UD-90521/2020\|EPI_ISL_671474\|2020- | ···················· | ···················· | ···················· | ···················· | ···················· | ···················· |
| T | 2786 hCoV-19/Hungary/UD-90554/2020\|EPI_ISL_671475\|2020- | ···················· | ···················· | ···················· | ···················· | ···················· | ···················· |
| T | 2787 hCoV-19/Hungary/UD-90625/2020\|EPI_ISL_671476\|2020- | ···················· | ···················· | ···················· | ···················· | ···················· | ···················· |
| T | 2788 hCoV-19/Hungary/UD-90635/2020\|EPI_ISL_671477\|2020- | ···················· | ···················· | ···················· | ···················· | ···················· | ···················· |

```
T   2789 hCoV-19/Hungary/UD-91370/2020|EPI_ISL_671478|2020-   ····················  ····················  ····················  ····················  ····················  ····················
T   2790 hCoV-19/Hungary/UD-91374/2020|EPI_ISL_671479|2020-   ····················  ····················  ····················  ····················  ····················  ····················
T   2791 hCoV-19/Hungary/UD-91459/2020|EPI_ISL_671480|2020-   ····················  ····················  ····················  ····················  ····················  ····················
T   2792 hCoV-19/Hungary/UD-91485/2020|EPI_ISL_671481|2020-   ····················  ····················  ····················  ····················  ····················  ····················
T   2793 hCoV-19/Hungary/UD-91486/2020|EPI_ISL_671482|2020-   ····················  ····················  ····················  ····················  ····················  ····················
T   2794 hCoV-19/Hungary/UD-91623/2020|EPI_ISL_671483|2020-   ····················  ····················  ····················  ····················  ····················  ····················
T   2795 hCoV-19/Hungary/UD-91823/2020|EPI_ISL_671484|2020-   ····················  ····················  ····················  ····················  ····················  ····················
T   2796 hCoV-19/Hungary/UD-93822/2020|EPI_ISL_671485|2020-   ····················  ····················  ····················  ····················  ····················  ····················
T   2797 hCoV-19/Canada/ON-PHL-20-02222/2020|EPI_ISL_671500   ····················  ····················  ····················  ····················  ····················  ····················
T   2798 hCoV-19/Canada/ON-PHL-20-02262/2020|EPI_ISL_671501   ····················  ····················  ····················  ····················  ····················  ····················
T   2799 hCoV-19/Canada/ON-PHL-20-02227/2020|EPI_ISL_671502   ····················  ····················  ····················  ····················  ····················  ····················
T   2800 hCoV-19/Canada/ON-PHL-20-02231/2020|EPI_ISL_671503   ····················  ····················  ····················  ····················  NNN·················  ····················   1
T   2802 hCoV-19/Canada/ON-PHL-20-02232/2020|EPI_ISL_671505   ····················  ····················  ····················  ····················  ····················  ····················
T   2803 hCoV-19/Canada/ON-PHL-20-02234/2020|EPI_ISL_671506   ····················  ····················  ····················  ····················  ·NN·················  ····················
T   2805 hCoV-19/Canada/ON-PHL-20-02209/2020|EPI_ISL_671508   ····················  ····················  ····················  ····················  ····················  ····················
T   2806 hCoV-19/Canada/ON-PHL-20-02213/2020|EPI_ISL_671509   ····················  ····················  ····················  ····················  ····················  ····················
T   2807 hCoV-19/Canada/ON-PHL-20-02239/2020|EPI_ISL_671510   ····················  ····················  ····················  ····················  ····················  ····················
T   2809 hCoV-19/Canada/ON-PHL-20-02241/2020|EPI_ISL_671512   ····················  ···A················  ····················  ····················  ····················  ····················
T   2810 hCoV-19/Canada/ON-PHL-20-02243/2020|EPI_ISL_671513   ····················  ····················  ····················  ····················  ····················  ····················
T   2811 hCoV-19/Canada/ON-PHL-20-02229/2020|EPI_ISL_671514   ····················  ····················  ····················  ····················  ····················  ····················
T   2812 hCoV-19/Canada/ON-PHL-20-02230/2020|EPI_ISL_671515   ····················  ····················  ····················  ····················  ····················  ····················
T   2813 hCoV-19/Canada/ON-PHL-20-02251/2020|EPI_ISL_671516   ····················  ····················  ····················  ····················  ····················  ····················
T   2814 hCoV-19/Canada/ON-PHL-20-02252/2020|EPI_ISL_671517   ····················  ····················  ····················  ····················  ····················  ····················
T   2815 hCoV-19/Canada/ON-PHL-20-02254/2020|EPI_ISL_671518   ····················  ····················  ····················  ····················  ····················  ····················
T   2817 hCoV-19/Belgium/ULG-10836/2020|EPI_ISL_666872|2020   ····················  ····················  ····················  ····················  ····················  ····················
T   2818 hCoV-19/Belgium/ULG-10842/2020|EPI_ISL_666877|2020   ····················  ····················  ····················  ····················  ····················  ····················
T   2819 hCoV-19/Belgium/ULG-10843/2020|EPI_ISL_666878|2020   ····················  ····················  ····················  ····················  ····················  ····················
T   2820 hCoV-19/Belgium/ULG-10844/2020|EPI_ISL_666879|2020   ····················  ····················  ····················  ····················  ····················  ····················
T   2821 hCoV-19/Belgium/ULG-10846/2020|EPI_ISL_666881|2020   ····················  ····················  ····················  ····················  ····················  ····················
T   2822 hCoV-19/Belgium/ULG-10847/2020|EPI_ISL_666882|2020   ····················  ····················  ····················  ····················  ····················  ····················
T   2823 hCoV-19/Belgium/ULG-10848/2020|EPI_ISL_666883|2020   ····················  ····················  ····················  ····················  ····················  ····················
T   2824 hCoV-19/Belgium/ULG-10849/2020|EPI_ISL_666884|2020   ····················  ····················  ····················  ····················  ····················  ····················
T   2825 hCoV-19/Belgium/ULG-10850/2020|EPI_ISL_666885|2020   ····················  ····················  ····················  ····················  ····················  ····················
T   2826 hCoV-19/Belgium/ULG-10851/2020|EPI_ISL_666886|2020   ····················  ····················  ····················  ····················  ····················  ····················
T   2827 hCoV-19/Belgium/ULG-10852/2020|EPI_ISL_666887|2020   ····················  ····················  ····················  ····················  ····················  ····················
T   2828 hCoV-19/USA/MI-MDHHS-SC22264/2020|EPI_ISL_666930|2   ····················  ····················  ····················  ····················  ····················  ····················
T   2829 hCoV-19/USA/MI-MDHHS-SC22265/2020|EPI_ISL_666931|2   ····················  ····················  ····················  ····················  ····················  ····················
T   2830 hCoV-19/USA/MI-MDHHS-SC22266/2020|EPI_ISL_666932|2   ····················  ····················  ····················  ····················  ····················  ····················
T   2831 hCoV-19/USA/MI-MDHHS-SC22267/2020|EPI_ISL_666933|2   ····················  ····················  ····················  ····················  ····················  ····················
T   2832 hCoV-19/USA/MI-MDHHS-SC22268/2020|EPI_ISL_666934|2   ····················  ····················  ····················  ····················  ····················  ····················
T   2833 hCoV-19/USA/MI-MDHHS-SC22269/2020|EPI_ISL_666935|2   ····················  ····················  ····················  ····················  ····················  ····················
T   2834 hCoV-19/USA/MI-MDHHS-SC22270/2020|EPI_ISL_666936|2   ····················  ·············A······  ····················  ····················  ····················  ····················
T   2835 hCoV-19/USA/MI-MDHHS-SC22271/2020|EPI_ISL_666937|2   ····················  ·············A······  ····················  ····················  ····················  ····················
T   2836 hCoV-19/USA/MI-MDHHS-SC22272/2020|EPI_ISL_666938|2   ····················  ····················  ····················  ····················  ····················  ····················
T   2837 hCoV-19/USA/MI-MDHHS-SC22273/2020|EPI_ISL_666939|2   ····················  ····················  ····················  ····················  ····················  ····················
T   2838 hCoV-19/USA/MI-MDHHS-SC22274/2020|EPI_ISL_666940|2   ····················  ····················  ····················  ····················  ····················  ····················
T   2839 hCoV-19/USA/MI-MDHHS-SC22275/2020|EPI_ISL_666941|2   ····················  ····················  ····················  ····················  ····················  ····················
T   2840 hCoV-19/USA/MI-MDHHS-SC22276/2020|EPI_ISL_666942|2   ····················  ····················  ····················  ····················  ····················  ····················
T   2841 hCoV-19/USA/MI-MDHHS-SC22277/2020|EPI_ISL_666943|2   ····················  ····················  ····················  ····················  ····················  ····················
T   2842 hCoV-19/USA/MI-MDHHS-SC22278/2020|EPI_ISL_666944|2   ····················  ····················  ····················  ····················  ····················  ····················
T   2843 hCoV-19/USA/MI-MDHHS-SC22279/2020|EPI_ISL_666945|2   ····················  ·············A······  ····················  ····················  ····················  ····················
T   2844 hCoV-19/USA/MI-MDHHS-SC22280/2020|EPI_ISL_666946|2   ····················  ····················  ····················  ····················  ····················  ····················
T   2845 hCoV-19/USA/MI-MDHHS-SC22281/2020|EPI_ISL_666947|2   ····················  ·A··················  ····················  ····················  ····················  ····················
T   2846 hCoV-19/USA/MI-MDHHS-SC22282/2020|EPI_ISL_666948|2   ····················  ····················  ····················  ····················  ····················  ····················
T   2847 hCoV-19/USA/MI-MDHHS-SC22283/2020|EPI_ISL_666949|2   ····················  ····················  ····················  ····················  ····················  ····················
T   2848 hCoV-19/USA/MI-MDHHS-SC22284/2020|EPI_ISL_666950|2   ····················  ····················  ····················  ····················  ····················  ····················
T   2849 hCoV-19/USA/MI-MDHHS-SC22285/2020|EPI_ISL_666951|2   ····················  ····················  ····················  ····················  ····················  ····················
T   2850 hCoV-19/USA/MI-MDHHS-SC22286/2020|EPI_ISL_666952|2   ····················  ····················  ····················  ····················  ····················  ····················
T   2851 hCoV-19/USA/MI-MDHHS-SC22287/2020|EPI_ISL_666953|2   ····················  ····················  ····················  ····················  ····················  ····················
T   2852 hCoV-19/USA/MI-MDHHS-SC22288/2020|EPI_ISL_666954|2   ····················  ·A··················  ····················  ····················  ····················  ····················
T   2853 hCoV-19/USA/MI-MDHHS-SC22289/2020|EPI_ISL_666955|2   ····················  ····················  ····················  ····················  ····················  ····················
T   2854 hCoV-19/USA/MI-MDHHS-SC22290/2020|EPI_ISL_666956|2   ····················  ····················  ····················  ····················  ····················  ····················
T   2855 hCoV-19/USA/MI-MDHHS-SC22291/2020|EPI_ISL_666957|2   ····················  ····················  ····················  ····················  ····················  ····················
T   2856 hCoV-19/USA/MI-MDHHS-SC22292/2020|EPI_ISL_666958|2   ····················  ····················  ····················  ····················  ····················  ····················
T   2857 hCoV-19/USA/MI-MDHHS-SC22293/2020|EPI_ISL_666959|2   ····················  ······A······A······  ····················  ····················  ····················  ····················
T   2858 hCoV-19/USA/MI-MDHHS-SC22294/2020|EPI_ISL_666960|2   ····················  ····················  ····················  ····················  ····················  ····················
T   2859 hCoV-19/USA/MI-MDHHS-SC22295/2020|EPI_ISL_666961|2   ····················  ·A··················  ····················  ····················  ····················  ····················
T   2860 hCoV-19/USA/MI-MDHHS-SC22296/2020|EPI_ISL_666962|2   ····················  ····················  ····················  ····················  ····················  ····················
T   2861 hCoV-19/USA/MI-MDHHS-SC22297/2020|EPI_ISL_666963|2   ····················  ·A··················  ····················  ····················  ····················  ····················
T   2862 hCoV-19/USA/NY-QDX-2748/2020|EPI_ISL_666964|2020-0   ····················  ····················  ····················  ····················  ····················  ····················
T   2863 hCoV-19/USA/CA-ALSR-4490/2020|EPI_ISL_666966|2020-   ····················  ····················  ····················  ····················  ····················  ····················
T   2864 hCoV-19/USA/CA-ALSR-4492/2020|EPI_ISL_666967|2020-   ····················  ····················  ····················  ····················  ····················  ····················
T   2865 hCoV-19/USA/CA-ALSR-4497/2020|EPI_ISL_666968|2020-   ····················  ·A··················  ····················  ····················  ····················  ····················
T   2866 hCoV-19/USA/CA-ALSR-4498/2020|EPI_ISL_666969|2020-   ····················  ····················  ····················  ····················  ····················  ····················
T   2867 hCoV-19/USA/CA-ALSR-4500/2020|EPI_ISL_666970|2020-   ····················  ····················  ····················  ····················  ····················  ····················
T   2868 hCoV-19/USA/CA-ALSR-4501/2020|EPI_ISL_666971|2020-   ····················  ····················  ····················  ····················  ····················  ····················
T   2869 hCoV-19/USA/CA-ALSR-4503/2020|EPI_ISL_666972|2020-   ····················  ····················  ····················  ····················  ····················  ····················
T   2870 hCoV-19/USA/CA-ALSR-4505/2020|EPI_ISL_666973|2020-   ····················  ·A··················  ····················  ····················  ····················  ····················
T   2871 hCoV-19/USA/CA-ALSR-4507/2020|EPI_ISL_666974|2020-   ····················  ····················  ····················  ····················  ····················  ····················
T   2872 hCoV-19/USA/CA-ALSR-4508/2020|EPI_ISL_666975|2020-   ····················  ····················  ····················  ····················  ····················  ····················
```

| | | | | | | | |
|---|---|---|---|---|---|---|---|
| T | 2873 hCoV-19/Denmark/DCGC-9671/2020\|EPI_ISL_669100\|2020 | .................... | .................... | .................... | .................... | .................... | .................... |
| T | 2874 hCoV-19/Denmark/DCGC-9672/2020\|EPI_ISL_669101\|2020 | .................... | .................... | .................... | .................... | .................... | .................... |
| T | 2875 hCoV-19/Denmark/DCGC-9673/2020\|EPI_ISL_669102\|2020 | .................... | .................... | .................... | .................... | .................... | .................... |
| T | 2876 hCoV-19/Denmark/DCGC-9674/2020\|EPI_ISL_669103\|2020 | .................... | .................... | .................... | .................... | .................... | .................... |
| T | 2877 hCoV-19/Denmark/DCGC-9676/2020\|EPI_ISL_669104\|2020 | .................... | .................... | .................... | .................... | .................... | .................... |
| T | 2878 hCoV-19/Denmark/DCGC-9677/2020\|EPI_ISL_669105\|2020 | .................... | .................... | .................... | .................... | .................... | .................... |
| T | 2879 hCoV-19/Denmark/DCGC-9679/2020\|EPI_ISL_669106\|2020 | .................... | .................... | .................... | .................... | .................... | .................... |
| T | 2880 hCoV-19/Denmark/DCGC-9680/2020\|EPI_ISL_669107\|2020 | .................... | .................... | .................... | .................... | .................... | .................... |
| T | 2881 hCoV-19/Denmark/DCGC-9682/2020\|EPI_ISL_669108\|2020 | .................... | .................... | .................... | .................... | .................... | .................... |
| T | 2882 hCoV-19/Denmark/DCGC-9686/2020\|EPI_ISL_669109\|2020 | .................... | .................... | .................... | .................... | .................... | .................... |
| T | 2883 hCoV-19/Denmark/DCGC-9688/2020\|EPI_ISL_669110\|2020 | .................... | .................... | .................... | .................... | .................... | .................... |
| T | 2884 hCoV-19/Denmark/DCGC-9689/2020\|EPI_ISL_669111\|2020 | .................... | .................... | .................... | .................... | .................... | .................... |
| T | 2885 hCoV-19/Denmark/DCGC-9690/2020\|EPI_ISL_669112\|2020 | .................... | .................... | .................... | .................... | .................... | .................... |
| T | 2886 hCoV-19/Denmark/DCGC-9691/2020\|EPI_ISL_669113\|2020 | .................... | .................... | .................... | .................... | .................... | .................... |
| T | 2887 hCoV-19/Denmark/DCGC-9692/2020\|EPI_ISL_669114\|2020 | .................... | .................... | .................... | .................... | .................... | .................... |
| T | 2888 hCoV-19/Denmark/DCGC-9696/2020\|EPI_ISL_669115\|2020 | .................... | .................... | .................... | .................... | .................... | .................... |
| T | 2889 hCoV-19/Denmark/DCGC-9697/2020\|EPI_ISL_669116\|2020 | .................... | .................... | .................... | .................... | .................... | .................... |
| T | 2890 hCoV-19/Denmark/DCGC-9698/2020\|EPI_ISL_669117\|2020 | .................... | .................... | .................... | .................... | .................... | .................... |
| T | 2891 hCoV-19/Denmark/DCGC-9701/2020\|EPI_ISL_669118\|2020 | .................... | .................... | .................... | .................... | .................... | .................... |
| T | 2892 hCoV-19/Denmark/DCGC-9703/2020\|EPI_ISL_669119\|2020 | .................... | .................... | .................... | .................... | .................... | .................... |
| T | 2893 hCoV-19/Denmark/DCGC-9706/2020\|EPI_ISL_669120\|2020 | .................... | .................... | .................... | .................... | .................... | .................... |
| T | 2894 hCoV-19/Denmark/DCGC-9709/2020\|EPI_ISL_669121\|2020 | .................... | .................... | .................... | .................... | .................... | .................... |
| T | 2895 hCoV-19/Denmark/DCGC-9710/2020\|EPI_ISL_669122\|2020 | .................... | .................... | .................... | .................... | .................... | .................... |
| T | 2896 hCoV-19/Denmark/DCGC-9714/2020\|EPI_ISL_669123\|2020 | .................... | .................... | .................... | .................... | .................... | .................... |
| T | 2897 hCoV-19/Denmark/DCGC-9715/2020\|EPI_ISL_669124\|2020 | .................... | .A.................. | .................... | .................... | .................... | .................... |
| T | 2898 hCoV-19/Denmark/DCGC-9716/2020\|EPI_ISL_669125\|2020 | .................... | .................... | .................... | .................... | .................... | .................... |
| T | 2899 hCoV-19/Denmark/DCGC-9719/2020\|EPI_ISL_669126\|2020 | .................... | .................... | .................... | .................... | .................... | .................... |
| T | 2900 hCoV-19/Denmark/DCGC-9720/2020\|EPI_ISL_669127\|2020 | .................... | .................... | .................... | .................... | .................... | .................... |
| T | 2901 hCoV-19/Denmark/DCGC-9722/2020\|EPI_ISL_669128\|2020 | .................... | .................... | .................... | .................... | .................... | .................... |
| T | 2902 hCoV-19/Denmark/DCGC-9725/2020\|EPI_ISL_669129\|2020 | .................... | .................... | .................... | .................... | .................... | .................... |
| T | 2903 hCoV-19/Denmark/DCGC-9726/2020\|EPI_ISL_669130\|2020 | .................... | .................... | .................... | .................... | .................... | .................... |
| T | 2904 hCoV-19/Denmark/DCGC-9727/2020\|EPI_ISL_669131\|2020 | .................... | .................... | .................... | .................... | .................... | .................... |
| T | 2905 hCoV-19/Denmark/DCGC-9730/2020\|EPI_ISL_669132\|2020 | .................... | .................... | .................... | .................... | .................... | .................... |
| T | 2906 hCoV-19/Denmark/DCGC-9731/2020\|EPI_ISL_669133\|2020 | .................... | ....A............... | .................... | .................... | .................... | .................... |
| T | 2907 hCoV-19/Denmark/DCGC-9733/2020\|EPI_ISL_669134\|2020 | .................... | .................... | .................... | .................... | .................... | .................... |
| T | 2908 hCoV-19/Denmark/DCGC-9734/2020\|EPI_ISL_669135\|2020 | .................... | .................... | .................... | .................... | .................... | .................... |
| T | 2909 hCoV-19/Denmark/DCGC-9737/2020\|EPI_ISL_669136\|2020 | .................... | .................... | .................... | .................... | .................... | .................... |
| T | 2910 hCoV-19/Denmark/DCGC-9738/2020\|EPI_ISL_669137\|2020 | .................... | .................... | .................... | .................... | .................... | .................... |
| T | 2911 hCoV-19/Denmark/DCGC-9740/2020\|EPI_ISL_669138\|2020 | .................... | .................... | .................... | .................... | .................... | .................... |
| T | 2912 hCoV-19/Denmark/DCGC-9742/2020\|EPI_ISL_669139\|2020 | .................... | .A.................. | .................... | .................... | .................... | ...A................ |
| T | 2913 hCoV-19/Denmark/DCGC-9743/2020\|EPI_ISL_669140\|2020 | .................... | .A.................. | .................... | .................... | .................... | .................... |
| T | 2914 hCoV-19/Denmark/DCGC-9745/2020\|EPI_ISL_669142\|2020 | .................... | .................... | .................... | .................... | .................... | .................... |
| T | 2915 hCoV-19/Denmark/DCGC-9746/2020\|EPI_ISL_669143\|2020 | .................... | .................... | .................... | .................... | .................... | .................... |
| T | 2916 hCoV-19/Denmark/DCGC-9747/2020\|EPI_ISL_669144\|2020 | .................... | .................... | .................... | .................... | .................... | .................... |
| T | 2917 hCoV-19/Denmark/DCGC-9748/2020\|EPI_ISL_669145\|2020 | .................... | .................... | .................... | .................... | .................... | .................... |
| T | 2918 hCoV-19/Denmark/DCGC-9749/2020\|EPI_ISL_669146\|2020 | .................... | ....A............... | .................... | .................... | .................... | .................... |
| T | 2919 hCoV-19/Denmark/DCGC-9750/2020\|EPI_ISL_669147\|2020 | .................... | .................... | .................... | .................... | .................... | .................... |
| T | 2920 hCoV-19/Denmark/DCGC-9751/2020\|EPI_ISL_669148\|2020 | .................... | .................... | .................... | .................... | .................... | .................... |
| T | 2921 hCoV-19/Denmark/DCGC-9753/2020\|EPI_ISL_669149\|2020 | .................... | .A.................. | .................... | .................... | .................... | .................... |
| T | 2922 hCoV-19/Denmark/DCGC-9778/2020\|EPI_ISL_669170\|2020 | .................... | .................... | .................... | .................... | .................... | .................... |
| T | 2923 hCoV-19/Denmark/DCGC-9779/2020\|EPI_ISL_669171\|2020 | .................... | .................... | .................... | .................... | .................... | .................... |
| T | 2924 hCoV-19/Denmark/DCGC-9780/2020\|EPI_ISL_669172\|2020 | .................... | .A.................. | .................... | .................... | .................... | .................... |
| T | 2925 hCoV-19/Denmark/DCGC-9781/2020\|EPI_ISL_669173\|2020 | .................... | .................... | .................... | .................... | .................... | .................... |
| T | 2926 hCoV-19/Denmark/DCGC-9782/2020\|EPI_ISL_669174\|2020 | .................... | .................... | .................... | .................... | .................... | .................... |
| T | 2927 hCoV-19/Denmark/DCGC-9784/2020\|EPI_ISL_669175\|2020 | .................... | .................... | .................... | .................... | .................... | .................... |
| T | 2928 hCoV-19/Denmark/DCGC-9785/2020\|EPI_ISL_669176\|2020 | .................... | .................... | .................... | .................... | .................... | .................... |
| T | 2929 hCoV-19/Denmark/DCGC-9788/2020\|EPI_ISL_669177\|2020 | .................... | .................... | .................... | .................... | .................... | .................... |
| T | 2930 hCoV-19/Denmark/DCGC-9789/2020\|EPI_ISL_669178\|2020 | .................... | .................... | .................... | .................... | .................... | .................... |
| T | 2931 hCoV-19/Denmark/DCGC-9790/2020\|EPI_ISL_669179\|2020 | .................... | .................... | .................... | .................... | .................... | .................... |
| T | 2932 hCoV-19/Denmark/DCGC-9791/2020\|EPI_ISL_669180\|2020 | .................... | .................... | .................... | .................... | .................... | .................... |
| T | 2933 hCoV-19/Denmark/DCGC-9792/2020\|EPI_ISL_669181\|2020 | .................... | .................... | .................... | .................... | .................... | .................... |
| T | 2934 hCoV-19/Denmark/DCGC-9793/2020\|EPI_ISL_669182\|2020 | .................... | .................... | .................... | .................... | .................... | .................... |
| T | 2935 hCoV-19/Denmark/DCGC-9794/2020\|EPI_ISL_669183\|2020 | .................... | .................... | .................... | .................... | .................... | .................... |
| T | 2936 hCoV-19/Denmark/DCGC-9795/2020\|EPI_ISL_669184\|2020 | .................... | .................... | .................... | .................... | .................... | .................... |
| T | 2937 hCoV-19/Denmark/DCGC-9796/2020\|EPI_ISL_669185\|2020 | .................... | .................... | .................... | .................... | .................... | .................... |
| T | 2938 hCoV-19/Denmark/DCGC-9797/2020\|EPI_ISL_669186\|2020 | .................... | .................... | .................... | .................... | .................... | .................... |
| T | 2939 hCoV-19/Denmark/DCGC-9798/2020\|EPI_ISL_669187\|2020 | .................... | .................... | .................... | .................... | .................... | .................... |
| T | 2940 hCoV-19/Denmark/DCGC-9799/2020\|EPI_ISL_669188\|2020 | .................... | .................... | .................... | .................... | .................... | .................... |
| T | 2941 hCoV-19/Denmark/DCGC-9800/2020\|EPI_ISL_669189\|2020 | .................... | .................... | .................... | .................... | .................... | .................... |
| T | 2942 hCoV-19/Denmark/DCGC-9801/2020\|EPI_ISL_669190\|2020 | .................... | .................... | .................... | .................... | .................... | .................... |
| T | 2943 hCoV-19/Denmark/DCGC-9802/2020\|EPI_ISL_669191\|2020 | .................... | .................... | .................... | .................... | .................... | .................... |
| T | 2944 hCoV-19/Denmark/DCGC-9803/2020\|EPI_ISL_669192\|2020 | .................... | .................... | .................... | .................... | .................... | .................... |
| T | 2945 hCoV-19/Denmark/DCGC-9804/2020\|EPI_ISL_669193\|2020 | .................... | .A.................. | .................... | .................... | .................... | .................... |
| T | 2946 hCoV-19/Denmark/DCGC-9806/2020\|EPI_ISL_669194\|2020 | .................... | .................... | .................... | .................... | .................... | .................... |
| T | 2947 hCoV-19/Denmark/DCGC-9807/2020\|EPI_ISL_669195\|2020 | .................... | .................... | .................... | .................... | .................... | .................... |
| T | 2948 hCoV-19/Denmark/DCGC-9808/2020\|EPI_ISL_669196\|2020 | .................... | .................... | .................... | .................... | .................... | .................... |
| T | 2949 hCoV-19/Denmark/DCGC-9809/2020\|EPI_ISL_669197\|2020 | .................... | .................... | .................... | .................... | .................... | .................... |
| T | 2950 hCoV-19/Denmark/DCGC-9810/2020\|EPI_ISL_669198\|2020 | .................... | .................... | .................... | .................... | .................... | .................... |
| T | 2951 hCoV-19/Denmark/DCGC-9813/2020\|EPI_ISL_669199\|2020 | .................... | .A.................. | .................... | .................... | .................... | .................... |
| T | 2952 hCoV-19/Denmark/DCGC-10626/2020\|EPI_ISL_670100\|202 | .................... | .................... | .................... | .................... | .................... | .................... |

| | | | | | | | | |
|---|---|---|---|---|---|---|---|---|
| T | 2953 hCoV-19/Denmark/DCGC-10627/2020\|EPI_ISL_670101\|202 | ···················· | ···················· | ····················· | ···················· | ···················· | ···················· | |
| T | 2954 hCoV-19/Denmark/DCGC-10628/2020\|EPI_ISL_670102\|202 | ···················· | ···················· | ····················· | ···················· | ···················· | ···················· | |
| T | 2955 hCoV-19/Denmark/DCGC-10629/2020\|EPI_ISL_670103\|202 | ···················· | ···················· | ····················· | ···················· | ···················· | ···················· | |
| T | 2956 hCoV-19/Denmark/DCGC-10631/2020\|EPI_ISL_670104\|202 | ···················· | ···················· | ····················· | ···················· | ···················· | ···················· | |
| T | 2957 hCoV-19/Denmark/DCGC-10633/2020\|EPI_ISL_670105\|202 | ···················· | ···················· | ····················· | ···················· | ···················· | ···················· | |
| T | 2958 hCoV-19/Denmark/DCGC-10636/2020\|EPI_ISL_670106\|202 | ···················· | A··················· | ····················· | ···················· | ···················· | ···················· | |
| T | 2959 hCoV-19/Denmark/DCGC-10637/2020\|EPI_ISL_670107\|202 | ···················· | ···················· | ····················· | ···················· | ···················· | ···················· | |
| T | 2960 hCoV-19/Denmark/DCGC-10639/2020\|EPI_ISL_670108\|202 | ···················· | ···················· | ····················· | ···················· | ···················· | ···················· | |
| T | 2961 hCoV-19/Denmark/DCGC-10641/2020\|EPI_ISL_670109\|202 | ···················· | ···················· | ····················· | ···················· | ···················· | ···················· | |
| T | 2962 hCoV-19/Denmark/DCGC-10643/2020\|EPI_ISL_670110\|202 | ···················· | ···················· | ····················· | ···················· | ···················· | ···················· | |
| T | 2963 hCoV-19/Denmark/DCGC-10646/2020\|EPI_ISL_670111\|202 | ···················· | ···················· | ····················· | ···················· | ···················· | ···················· | |
| T | 2964 hCoV-19/Denmark/DCGC-10647/2020\|EPI_ISL_670112\|202 | ···················· | ···················· | ····················· | ···················· | ···················· | ···················· | |
| T | 2965 hCoV-19/Denmark/DCGC-10648/2020\|EPI_ISL_670113\|202 | ···················· | ···················· | ····················· | ···················· | ···················· | ···················· | |
| T | 2966 hCoV-19/Denmark/DCGC-10649/2020\|EPI_ISL_670114\|202 | ···················· | ···················· | ····················· | ···················· | ···················· | ···················· | |
| T | 2967 hCoV-19/Denmark/DCGC-10650/2020\|EPI_ISL_670115\|202 | ···················· | ·A·················· | ····················· | ···················· | ···················· | ··A················· | |
| T | 2968 hCoV-19/Denmark/DCGC-10651/2020\|EPI_ISL_670116\|202 | ···················· | ···················· | ····················· | ···················· | ···················· | ···················· | |
| T | 2969 hCoV-19/Denmark/DCGC-10652/2020\|EPI_ISL_670117\|202 | ···················· | ···················· | ····················· | ···················· | ···················· | ···················· | |
| T | 2970 hCoV-19/Denmark/DCGC-10653/2020\|EPI_ISL_670118\|202 | ···················· | ···················· | ····················· | ···················· | ···················· | ···················· | |
| T | 2971 hCoV-19/Denmark/DCGC-10656/2020\|EPI_ISL_670119\|202 | ···················· | ···················· | ····················· | ···················· | ···················· | ···················· | |
| T | 2972 hCoV-19/Denmark/DCGC-10658/2020\|EPI_ISL_670120\|202 | ···················· | ···················· | ····················· | ···················· | ···················· | ···················· | |
| T | 2973 hCoV-19/Denmark/DCGC-10661/2020\|EPI_ISL_670121\|202 | ···················· | ···················· | ····················· | ···················· | ···················· | ···················· | |
| T | 2974 hCoV-19/Denmark/DCGC-10679/2020\|EPI_ISL_670131\|202 | ···················· | ···················· | ····················· | ···················· | ···················· | ···················· | |
| T | 2975 hCoV-19/Denmark/DCGC-10680/2020\|EPI_ISL_670132\|202 | ···················· | ···················· | ····················· | ···················· | ···················· | ···················· | |
| T | 2976 hCoV-19/Denmark/DCGC-10712/2020\|EPI_ISL_670155\|202 | ···················· | ···················· | ····················· | ···················· | ···················· | ···················· | |
| T | 2977 hCoV-19/Denmark/DCGC-10713/2020\|EPI_ISL_670156\|202 | ···················· | ···················· | ····················· | ···················· | ···················· | ···················· | |
| T | 2978 hCoV-19/Denmark/DCGC-10714/2020\|EPI_ISL_670157\|202 | ···················· | ···················· | ····················· | ···················· | ···················· | ···················· | |
| T | 2979 hCoV-19/Denmark/DCGC-10715/2020\|EPI_ISL_670158\|202 | ···················· | ···················· | ····················· | ···················· | ···················· | ···················· | |
| T | 2980 hCoV-19/Denmark/DCGC-10717/2020\|EPI_ISL_670159\|202 | ···················· | ···················· | ····················· | ···················· | ···················· | ···················· | |
| T | 2981 hCoV-19/Denmark/DCGC-10718/2020\|EPI_ISL_670160\|202 | ···················· | ···················· | ····················· | ···················· | ···················· | ···················· | |
| T | 2982 hCoV-19/Denmark/DCGC-10719/2020\|EPI_ISL_670161\|202 | ···················· | ···················· | ····················· | ···················· | ···················· | ···················· | |
| T | 2983 hCoV-19/Denmark/DCGC-10722/2020\|EPI_ISL_670162\|202 | ···················· | ···················· | ····················· | ···················· | ···················· | ···················· | |
| T | 2984 hCoV-19/Denmark/DCGC-10725/2020\|EPI_ISL_670163\|202 | ···················· | ···················· | ····················· | ···················· | ···················· | ···················· | |
| T | 2985 hCoV-19/Denmark/DCGC-10726/2020\|EPI_ISL_670164\|202 | ···················· | ···················· | ····················· | ···················· | ···················· | ···················· | |
| T | 2986 hCoV-19/Denmark/DCGC-10728/2020\|EPI_ISL_670165\|202 | ···················· | ···················· | ····················· | ···················· | ···················· | ···················· | |
| T | 2987 hCoV-19/Denmark/DCGC-10729/2020\|EPI_ISL_670166\|202 | ···················· | ···················· | ····················· | ···················· | ···················· | ···················· | |
| T | 2988 hCoV-19/Denmark/DCGC-10730/2020\|EPI_ISL_670167\|202 | ···················· | ·A·················· | ····················· | ···················· | ···················· | ···················· | |
| T | 2989 hCoV-19/Denmark/DCGC-10731/2020\|EPI_ISL_670168\|202 | ···················· | ···················· | ····················· | ···················· | ···················· | ···················· | |
| T | 2990 hCoV-19/Denmark/DCGC-10734/2020\|EPI_ISL_670169\|202 | ···················· | ···················· | ····················· | ···················· | ···················· | ···················· | |
| T | 2991 hCoV-19/Denmark/DCGC-10735/2020\|EPI_ISL_670170\|202 | ···················· | ···················· | ····················· | ···················· | ···················· | ···················· | |
| T | 2992 hCoV-19/Denmark/DCGC-10737/2020\|EPI_ISL_670171\|202 | ···················· | ···················· | ····················· | ···················· | ···················· | ···················· | |
| T | 2993 hCoV-19/Denmark/DCGC-10738/2020\|EPI_ISL_670172\|202 | ···················· | ···················· | ····················· | ···················· | ···················· | ···················· | |
| T | 2994 hCoV-19/Denmark/DCGC-10739/2020\|EPI_ISL_670173\|202 | ···················· | ···················· | ····················· | ···················· | ···················· | ···················· | |
| T | 2995 hCoV-19/Denmark/DCGC-10740/2020\|EPI_ISL_670174\|202 | ···················· | ·A·················· | ····················· | ···················· | ···················· | ··A················· | |
| T | 2996 hCoV-19/Denmark/DCGC-10742/2020\|EPI_ISL_670175\|202 | ···················· | ···················· | ····················· | ···················· | ···················· | ···················· | |
| T | 2997 hCoV-19/Denmark/DCGC-10743/2020\|EPI_ISL_670176\|202 | ···················· | ···················· | ····················· | ···················· | ···················· | ···················· | |
| T | 2998 hCoV-19/Denmark/DCGC-10744/2020\|EPI_ISL_670177\|202 | ···················· | ···················· | ····················· | ···················· | ···················· | ···················· | |
| T | 2999 hCoV-19/Denmark/DCGC-10745/2020\|EPI_ISL_670178\|202 | ···················· | ···················· | ····················· | ···················· | ···················· | ···················· | |
| T | 3000 hCoV-19/Denmark/DCGC-10746/2020\|EPI_ISL_670179\|202 | ···················· | ···················· | ····················· | ···················· | ···················· | ···················· | |
| T | 3001 hCoV-19/Denmark/DCGC-10747/2020\|EPI_ISL_670180\|202 | ···················· | ···················· | ····················· | ···················· | ···················· | ···················· | |
| T | 3002 hCoV-19/Denmark/DCGC-10748/2020\|EPI_ISL_670181\|202 | ···················· | ···················· | ····················· | ···················· | ···················· | ···················· | |
| T | 3003 hCoV-19/Denmark/DCGC-10750/2020\|EPI_ISL_670182\|202 | ···················· | ···················· | ····················· | ···················· | ···················· | ···················· | |
| T | 3004 hCoV-19/Denmark/DCGC-10751/2020\|EPI_ISL_670183\|202 | ···················· | ···················· | ····················· | ···················· | ···················· | ···················· | |
| T | 3005 hCoV-19/Denmark/DCGC-10753/2020\|EPI_ISL_670184\|202 | ···················· | ···················· | ····················· | ···················· | ···················· | ···················· | |
| T | 3006 hCoV-19/Denmark/DCGC-10754/2020\|EPI_ISL_670185\|202 | ···················· | ···················· | ····················· | ···················· | ···················· | ···················· | |
| T | 3007 hCoV-19/Denmark/DCGC-10755/2020\|EPI_ISL_670186\|202 | ···················· | ···················· | ····················· | ···················· | ···················· | ···················· | |
| T | 3008 hCoV-19/Denmark/DCGC-10756/2020\|EPI_ISL_670187\|202 | ···················· | ·A·················· | ····················· | ···················· | ···················· | ···················· | |
| T | 3009 hCoV-19/Denmark/DCGC-10759/2020\|EPI_ISL_670188\|202 | ···················· | ···················· | ····················· | ···················· | ···················· | ···················· | |
| T | 3010 hCoV-19/Denmark/DCGC-10761/2020\|EPI_ISL_670189\|202 | ···················· | ·········A·········· | ····················· | ···················· | ···················· | ···················· | |
| T | 3011 hCoV-19/Denmark/DCGC-10762/2020\|EPI_ISL_670190\|202 | ···················· | ···················· | ····················· | ···················· | ···················· | ···················· | |
| T | 3012 hCoV-19/Denmark/DCGC-10763/2020\|EPI_ISL_670191\|202 | ···················· | ···················· | ····················· | ···················· | ···················· | ···················· | |
| T | 3013 hCoV-19/Denmark/DCGC-10764/2020\|EPI_ISL_670192\|202 | ···················· | ···················· | ····················· | ···················· | ···················· | ···················· | |
| T | 3014 hCoV-19/Denmark/DCGC-10765/2020\|EPI_ISL_670193\|202 | ···················· | ···················· | ····················· | ···················· | ···················· | ···················· | |
| T | 3015 hCoV-19/Denmark/DCGC-10766/2020\|EPI_ISL_670194\|202 | ···················· | ···················· | ····················· | ···················· | ···················· | ···················· | |
| T | 3016 hCoV-19/Denmark/DCGC-10767/2020\|EPI_ISL_670195\|202 | ···················· | ···················· | ····················· | ···················· | ···················· | ···················· | |
| T | 3017 hCoV-19/Denmark/DCGC-10768/2020\|EPI_ISL_670196\|202 | ···················· | ···················· | ····················· | ···················· | ···················· | ···················· | |
| T | 3018 hCoV-19/Denmark/DCGC-10769/2020\|EPI_ISL_670197\|202 | ···················· | ···················· | ····················· | ···················· | ···················· | ···················· | |
| T | 3019 hCoV-19/Denmark/DCGC-10772/2020\|EPI_ISL_670198\|202 | ···················· | ···················· | ····················· | ···················· | ···················· | ···················· | |
| T | 3020 hCoV-19/Denmark/DCGC-10773/2020\|EPI_ISL_670199\|202 | ···················· | ···················· | ····················· | ···················· | ···················· | ···················· | |
| T | 3021 hCoV-19/Ireland/LS-NVRL-20Z06652/2020\|EPI_ISL_6714 | ···················· | ···················· | ····················· | ···················· | ···················· | ···················· | |
| T | 3022 hCoV-19/Ireland/D-NVRL-20Z06241/2020\|EPI_ISL_67140 | ···················· | ···················· | ····················· | ···················· | ···················· | ···················· | |
| T | 3023 hCoV-19/Ireland/MN-NVRL-20Z06659/2020\|EPI_ISL_6714 | ···················· | ···················· | ····················· | ···················· | ···················· | ···················· | |
| T | 3024 hCoV-19/Hungary/SRC-1014/2020\|EPI_ISL_671414\|2020- | ···················· | ···················· | ····················· | ···················· | ···················· | ···················· | |
| T | 3025 hCoV-19/Hungary/SRC-1650/2020\|EPI_ISL_671415\|2020- | ···················· | ···················· | ····················· | ···················· | ···················· | ···················· | |
| T | 3026 hCoV-19/Hungary/SRC-2135/2020\|EPI_ISL_671416\|2020- | ···················· | ···················· | ····················· | ···················· | ···················· | ···················· | |
| T | 3028 hCoV-19/Hungary/SRC-2145/2020\|EPI_ISL_671418\|2020- | ···················· | ···················· | ····················· | ···················· | ···················· | ···················· | |
| T | 3029 hCoV-19/Hungary/UD-55426/2020\|EPI_ISL_671419\|2020- | ···················· | ··················C· | ····················· | ···················· | ···················· | ···················· | 1 |
| T | 3030 hCoV-19/Hungary/UD-55641/2020\|EPI_ISL_671420\|2020- | ···················· | ···················· | ····················· | ···················· | ···················· | ···················· | |
| T | 3031 hCoV-19/Hungary/UD-56251/2020\|EPI_ISL_671421\|2020- | ···················· | ···················· | ····················· | ···················· | ···················· | ···················· | |
| T | 3032 hCoV-19/Hungary/UD-56279/2020\|EPI_ISL_671423\|2020- | ···················· | ···················· | ····················· | ···················· | ···················· | ···················· | |
| T | 3033 hCoV-19/Hungary/UD-56284/2020\|EPI_ISL_671424\|2020- | ···················· | ···················· | ····················· | ···················· | ···················· | ···················· | |

T 3034 hCoV-19/Hungary/UD-56297/2020|EPI_ISL_671425|2020- ···················· ···················· ···················· ···················· ···················· ····················
T 3035 hCoV-19/Hungary/UD-56311/2020|EPI_ISL_671427|2020- ···················· ···················· ···················· ···················· ···················· ····················
T 3036 hCoV-19/Hungary/UD-56412/2020|EPI_ISL_671428|2020- ···················· ···················· ···················· ···················· ···················· ····················
T 3037 hCoV-19/Hungary/UD-56569/2020|EPI_ISL_671429|2020- ···················· ···················· ···················· ···················· ···················· ····················
T 3038 hCoV-19/Hungary/UD-56626/2020|EPI_ISL_671430|2020- ···················· ···················· ···················· ···················· ···················· ····················
T 3039 hCoV-19/Hungary/UD-57410/2020|EPI_ISL_671431|2020- ···················· ···················· ···················· ···················· ···················· ····················
T 3040 hCoV-19/Hungary/UD-57411/2020|EPI_ISL_671432|2020- ···················· ···················· ···················· ···················· ···················· ····················
T 3041 hCoV-19/Hungary/UD-57837/2020|EPI_ISL_671433|2020- ···················· ···················· ···················· ···················· ···················· ····················
T 3042 hCoV-19/Hungary/UD-57884/2020|EPI_ISL_671434|2020- ···················· ···················· ···················· ···················· ···················· ····················
T 3043 hCoV-19/Hungary/UD-57885/2020|EPI_ISL_671435|2020- ···················· ···················· ···················· ···················· ···················· ····················
T 3044 hCoV-19/Hungary/UD-57889/2020|EPI_ISL_671436|2020- ···················· ···················· ···················· ···················· ···················· ····················
T 3045 hCoV-19/Hungary/UD-57949/2020|EPI_ISL_671437|2020- ···················· ···················· ···················· ···················· ···················· ····················
T 3046 hCoV-19/Hungary/UD-57951/2020|EPI_ISL_671438|2020- ···················· ···················· ···················· ···················· ···················· ····················
T 3047 hCoV-19/Hungary/UD-57958/2020|EPI_ISL_671440|2020- ···················· ···················· ···················· ···················· ···················· ····················
T 3048 hCoV-19/Hungary/UD-57964/2020|EPI_ISL_671441|2020- ···················· ···················· ···················· ···················· ···················· ····················
T 3049 hCoV-19/Hungary/UD-57974/2020|EPI_ISL_671443|2020- ···················· ···················· ···················· ···················· ···················· ····················
T 3050 hCoV-19/Hungary/UD-57983/2020|EPI_ISL_671444|2020- ···················· ···················· ···················· ···················· ···················· ····················
T 3051 hCoV-19/Hungary/UD-57985/2020|EPI_ISL_671445|2020- ···················· ···················· ···················· ···················· ···················· ····················
T 3052 hCoV-19/Hungary/UD-58004/2020|EPI_ISL_671447|2020- ···················· ···················· ···················· ···················· ···················· ····················
T 3053 hCoV-19/Hungary/UD-58011/2020|EPI_ISL_671448|2020- ···················· ···················· ···················· ···················· ···················· ····················
T 3054 hCoV-19/Hungary/UD-58062/2020|EPI_ISL_671449|2020- ···················· ···················· ···················· ···················· ···················· ····················
T 3055 hCoV-19/Hungary/UD-58207/2020|EPI_ISL_671451|2020- ···················· ···················· ···················· ···················· ···················· ···············A····
T 3056 hCoV-19/Hungary/UD-58213/2020|EPI_ISL_671452|2020- ···················· ···················· ···················· ···················· ···················· ····················
T 3057 hCoV-19/Hungary/UD-90323/2020|EPI_ISL_671460|2020- ···················· ···················· ···················· ···················· ···················· ····················
T 3058 hCoV-19/Hungary/UD-90341/2020|EPI_ISL_671461|2020- ···················· ···················· ···················· ···················· ···················· ····················
T 3060 hCoV-19/Hungary/UD-93827/2020|EPI_ISL_671487|2020- ···················· ···················· ···················· ···················· ···················· ····················
T 3061 hCoV-19/Hungary/UD-93941/2020|EPI_ISL_671488|2020- ···················· ···················· ···················· ···················· ···················· ····················
T 3062 hCoV-19/Hungary/UD-93943/2020|EPI_ISL_671489|2020- ···················· ···················· ···················· ···················· ···················· ····················
T 3063 hCoV-19/Hungary/UD-93945/2020|EPI_ISL_671490|2020- ···················· ···················· ···················· ···················· ···················· ····················
T 3064 hCoV-19/Hungary/UD-93992/2020|EPI_ISL_671491|2020- ···················· ···················· ···················· ···················· ···················· ····················
T 3065 hCoV-19/Hungary/UD-93998/2020|EPI_ISL_671492|2020- ···················· ···················· ···················· ···················· ···················· ····················
T 3066 hCoV-19/Hungary/UD-94020/2020|EPI_ISL_671493|2020- ···················· ···················· ···················· ···················· ···················· ····················
T 3067 hCoV-19/Hungary/UD-94053/2020|EPI_ISL_671494|2020- ···················· ···················· ···················· ···················· ···················· ····················
T 3068 hCoV-19/Hungary/UD-93928/2020|EPI_ISL_671495|2020- ···················· ···················· ···················· ···················· ···················· ····················
T 3069 hCoV-19/Canada/ON-PHL-20-02196/2020|EPI_ISL_671496 ···················· ···················· ···················· ···················· ···················· ····················
T 3071 hCoV-19/Canada/ON-PHL-20-02258/2020|EPI_ISL_671498 ···················· ···················· ···················· ···················· ···················· ····················
T 3072 hCoV-19/Canada/ON-PHL-20-02259/2020|EPI_ISL_671499 ···················· ···················· ···················· ···················· ···················· ····················
T 3073 hCoV-19/England/QEUH-B673E5/2020|EPI_ISL_661960|20 ···················· ···················· ···················· ···················· ···················· ····················
T 3074 hCoV-19/England/ALDP-B7438B/2020|EPI_ISL_661961|20 ···················· ···················· ···················· ···················· ···················· ····················
T 3075 hCoV-19/England/QEUH-B681AE/2020|EPI_ISL_661963|20 ···················· ···················· ···················· ···················· ···················· ····················
T 3076 hCoV-19/England/ALDP-B7437C/2020|EPI_ISL_661964|20 ···················· ···················· ···················· ···················· ···················· ····················
T 3077 hCoV-19/England/ALDP-B74A0E/2020|EPI_ISL_661965|20 ···················· ···················· ···················· ···················· ···················· ····················
T 3078 hCoV-19/England/ALDP-B74075/2020|EPI_ISL_661966|20 ···················· ···················· ···················· ···················· ···················· ····················
T 3079 hCoV-19/England/QEUH-B6893D/2020|EPI_ISL_661967|20 ···················· ···················· ···················· ···················· ·····T·············· ····················
T 3080 hCoV-19/England/ALDP-B74CCC/2020|EPI_ISL_661968|20 ···················· ···················· ···················· ···················· ···················· ····················
T 3081 hCoV-19/England/ALDP-B755B1/2020|EPI_ISL_661970|20 ···················· ···················· ···················· ···················· ···················· ····················
T 3082 hCoV-19/England/QEUH-B68D04/2020|EPI_ISL_661971|20 ···················· ···················· ···················· ···················· ···················· ····················
T 3083 hCoV-19/England/QEUH-B686CD/2020|EPI_ISL_661972|20 ···················· ···················· ···················· ···················· ···················· ····················
T 3084 hCoV-19/England/QEUH-B68D31/2020|EPI_ISL_661973|20 ···················· ···················· ···················· ···················· ···················· ····················
T 3085 hCoV-19/England/ALDP-B75162/2020|EPI_ISL_661974|20 ···················· ···················· ···················· ···················· ···················· ····················
T 3086 hCoV-19/England/QEUH-B69170/2020|EPI_ISL_661975|20 ···················· ···················· ···················· ···················· ···················· ····················
T 3087 hCoV-19/England/QEUH-B68F86/2020|EPI_ISL_661976|20 ···················· ···················· ···················· ···················· ···················· ····················
T 3088 hCoV-19/England/ALDP-B75083/2020|EPI_ISL_661977|20 ···················· ···················· ···················· ···················· ···················· ····················
T 3089 hCoV-19/England/ALDP-B74998/2020|EPI_ISL_661978|20 ···················· ···················· ···················· ···················· ···················· ····················
T 3090 hCoV-19/England/QEUH-B6928C/2020|EPI_ISL_661979|20 ···················· ···················· ···················· ···················· ···················· ····················
T 3091 hCoV-19/England/ALDP-B740B1/2020|EPI_ISL_661980|20 ···················· ···················· ···················· ···················· ···················· ····················
T 3092 hCoV-19/England/ALDP-B74CAE/2020|EPI_ISL_661982|20 ···················· ···················· ···················· ···················· ···················· ····················
T 3093 hCoV-19/England/ALDP-B743A9/2020|EPI_ISL_661983|20 ···················· ···················· ···················· ···················· ···················· ····················
T 3094 hCoV-19/England/QEUH-B67FA5/2020|EPI_ISL_661984|20 ···················· ···················· ···················· ···················· ···················· ····················
T 3095 hCoV-19/England/ALDP-B749B6/2020|EPI_ISL_661985|20 ···················· ···················· ···················· ···················· ···················· ····················
T 3096 hCoV-19/England/QEUH-B67400/2020|EPI_ISL_661987|20 ···················· ···················· ···················· ···················· ···················· ····················
T 3097 hCoV-19/England/ALDP-B746EC/2020|EPI_ISL_661988|20 ···················· ···················· ···················· ···················· ···················· ····················
T 3098 hCoV-19/England/ALDP-B7495C/2020|EPI_ISL_661989|20 ···················· ···················· ···················· ···················· ···················· ····················
T 3099 hCoV-19/England/ALDP-B7500B/2020|EPI_ISL_661990|20 ···················· ···················· ···················· ···················· ···················· ····················
T 3100 hCoV-19/England/ALDP-B755A2/2020|EPI_ISL_661991|20 ···················· ···················· ···················· ···················· ···················· ····················
T 3101 hCoV-19/England/ALDP-B75302/2020|EPI_ISL_661992|20 ···················· ···················· ···················· ···················· ···················· ····················
T 3102 hCoV-19/England/QEUH-B68ED4/2020|EPI_ISL_661993|20 ···················· ·A·················· ···················· ···················· ···················· ··A·················
T 3103 hCoV-19/England/QEUH-B68E10/2020|EPI_ISL_661994|20 ···················· ···················· ···················· ···················· ···················· ····················
T 3104 hCoV-19/England/ALDP-B75171/2020|EPI_ISL_661995|20 ···················· ···················· ···················· ···················· ···················· ····················
T 3105 hCoV-19/England/QEUH-B68690/2020|EPI_ISL_661996|20 ···················· ···················· ···················· ···················· ·····T·············· ····················
T 3106 hCoV-19/England/QEUH-B67F3C/2020|EPI_ISL_661997|20 ···················· ···················· ···················· ···················· ···················· ····················
T 3107 hCoV-19/England/ALDP-B74DC9/2020|EPI_ISL_661998|20 ···················· ···················· ···················· ···················· ···················· ····················
T 3108 hCoV-19/England/ALDP-B7529C/2020|EPI_ISL_661999|20 ···················· ···················· ···················· ···················· ···················· ····················
T 3109 hCoV-19/USA/MD-MDH-0432/2020|EPI_ISL_666800|2020-1 ···················· ···················· ···················· ···················· ···················· ····················
T 3110 hCoV-19/USA/MD-MDH-0433/2020|EPI_ISL_666801|2020-1 ···················· ···················· ···················· ···················· ···················· ····················
T 3111 hCoV-19/USA/MD-MDH-0434/2020|EPI_ISL_666802|2020-1 ···················· ···················· ···················· ···················· ···················· ····················
T 3112 hCoV-19/USA/MD-MDH-0435/2020|EPI_ISL_666803|2020-1 ···················· ···················· ···················· ···················· ···················· ····················
T 3113 hCoV-19/USA/MD-MDH-0437/2020|EPI_ISL_666804|2020-1 ···················· ·A·················· ···················· ···················· ···················· ····················
T 3114 hCoV-19/USA/MD-MDH-0438/2020|EPI_ISL_666805|2020-1 ···················· ···················· ···················· ···················· ···················· ····················
T 3115 hCoV-19/USA/MD-MDH-0439/2020|EPI_ISL_666806|2020-1 ···················· ·A·················· ···················· ···················· ···················· ····················

| | | | | | | | |
|---|---|---|---|---|---|---|---|
| T | 3116 hCoV-19/USA/MD-MDH-0440/2020\|EPI_ISL_666807\|2020-1 | ···················· | ···················· | ····················· | ···················· | ···················· | ···················· |
| T | 3117 hCoV-19/USA/MD-MDH-0441/2020\|EPI_ISL_666808\|2020-1 | ···················· | ·A·················· | ····················· | ···················· | ···················· | ···················· |
| T | 3118 hCoV-19/USA/MD-MDH-0442/2020\|EPI_ISL_666809\|2020-1 | ···················· | ·A·················· | ····················· | ···················· | ···················· | ···················· |
| T | 3119 hCoV-19/USA/MD-MDH-0444/2020\|EPI_ISL_666811\|2020-1 | ···················· | ···················· | ····················· | ···················· | ···················· | ···················· |
| T | 3120 hCoV-19/USA/MD-MDH-0445/2020\|EPI_ISL_666812\|2020-1 | ···················· | ···················· | ····················· | ···················· | ···················· | ···················· |
| T | 3121 hCoV-19/USA/MD-MDH-0446/2020\|EPI_ISL_666813\|2020-1 | ···················· | ···················· | ····················· | ···················· | ···················· | ···················· |
| T | 3122 hCoV-19/USA/FL-BPHL-2203/2020\|EPI_ISL_666825\|2020- | ···················· | ···················· | ····················· | ···················· | ···················· | ···················· |
| T | 3123 hCoV-19/USA/FL-BPHL-2211/2020\|EPI_ISL_666833\|2020- | ···················· | ···················· | ····················· | ···················· | ···················· | ···················· |
| T | 3124 hCoV-19/USA/FL-BPHL-2214/2020\|EPI_ISL_666836\|2020- | ···················· | ···················· | ····················· | ···················· | ···················· | ···················· |
| T | 3128 hCoV-19/USA/FL-BPHL-2225/2020\|EPI_ISL_666847\|2020- | ···················· | ···················· | ····················· | ···················· | ···················· | ···················· |
| T | 3129 hCoV-19/USA/FL-BPHL-2227/2020\|EPI_ISL_666849\|2020- | ···················· | ···················· | ····················· | ···················· | ···················· | ···················· |
| T | 3130 hCoV-19/USA/FL-BPHL-2230/2020\|EPI_ISL_666852\|2020- | ···················· | ···A················ | ····················· | ···················· | ···················· | ···················· |
| T | 3131 hCoV-19/USA/FL-BPHL-2232/2020\|EPI_ISL_666854\|2020- | ···················· | ···················· | ····················· | ···················· | ···················· | ···················· |
| T | 3132 hCoV-19/France/NOR-UCN19/2020\|EPI_ISL_666870\|2020- | ···················· | ···················· | ····················· | ···················· | ···················· | ···················· |
| T | 3133 hCoV-19/Belgium/ULG-10838/2020\|EPI_ISL_666873\|2020 | ···················· | ·A·················· | ····················· | ···················· | ···················· | ···················· |
| T | 3134 hCoV-19/Belgium/ULG-10839/2020\|EPI_ISL_666874\|2020 | ···················· | ···················· | ····················· | ···················· | ···················· | ···················· |
| T | 3135 hCoV-19/Denmark/DCGC-9492/2020\|EPI_ISL_669000\|2020 | ···················· | ···················· | ····················· | ···················· | ···················· | ···················· |
| T | 3136 hCoV-19/Denmark/DCGC-9494/2020\|EPI_ISL_669001\|2020 | ···················· | ···················· | ····················· | ···················· | ···················· | ···················· |
| T | 3137 hCoV-19/Denmark/DCGC-9496/2020\|EPI_ISL_669002\|2020 | ···················· | ···················· | ····················· | ···················· | ···················· | ···················· |
| T | 3138 hCoV-19/Denmark/DCGC-9499/2020\|EPI_ISL_669003\|2020 | ···················· | ···················· | ····················· | ···················· | ···················· | ···················· |
| T | 3139 hCoV-19/Denmark/DCGC-9500/2020\|EPI_ISL_669004\|2020 | ···················· | ···················· | ····················· | ···················· | ···················· | ···················· |
| T | 3140 hCoV-19/Denmark/DCGC-9502/2020\|EPI_ISL_669005\|2020 | ···················· | ···················· | ····················· | ···················· | ···················· | ···················· |
| T | 3141 hCoV-19/Denmark/DCGC-9503/2020\|EPI_ISL_669006\|2020 | ···················· | ···················· | ····················· | ···················· | ···················· | ···················· |
| T | 3142 hCoV-19/Denmark/DCGC-9505/2020\|EPI_ISL_669007\|2020 | ···················· | ···················· | ····················· | ···················· | ···················· | ···················· |
| T | 3143 hCoV-19/Denmark/DCGC-9506/2020\|EPI_ISL_669008\|2020 | ···················· | ···················· | ····················· | ···················· | ···················· | ···················· |
| T | 3144 hCoV-19/Denmark/DCGC-9507/2020\|EPI_ISL_669009\|2020 | ···················· | ···················· | ····················· | ···················· | ···················· | ···················· |
| T | 3145 hCoV-19/Denmark/DCGC-9508/2020\|EPI_ISL_669010\|2020 | ···················· | ···················· | ····················· | ···················· | ···················· | ···················· |
| T | 3146 hCoV-19/Denmark/DCGC-9510/2020\|EPI_ISL_669011\|2020 | ···················· | ···················· | ····················· | ···················· | ···················· | ···················· |
| T | 3147 hCoV-19/Denmark/DCGC-9511/2020\|EPI_ISL_669012\|2020 | ···················· | ·A·················· | ····················· | ···················· | ···················· | ··A················· |
| T | 3148 hCoV-19/Denmark/DCGC-9513/2020\|EPI_ISL_669013\|2020 | ···················· | ···················· | ····················· | ···················· | ···················· | ···················· |
| T | 3149 hCoV-19/Denmark/DCGC-9514/2020\|EPI_ISL_669014\|2020 | ···················· | ···················· | ····················· | ···················· | ···················· | ···················· |
| T | 3150 hCoV-19/Denmark/DCGC-9517/2020\|EPI_ISL_669015\|2020 | ···················· | ···················· | ····················· | ···················· | ···················· | ···················· |
| T | 3151 hCoV-19/Denmark/DCGC-9519/2020\|EPI_ISL_669016\|2020 | ···················· | ···················· | ····················· | ···················· | ···················· | ···················· |
| T | 3152 hCoV-19/Denmark/DCGC-9520/2020\|EPI_ISL_669017\|2020 | ···················· | ···················· | ····················· | ···················· | ···················· | ···················· |
| T | 3153 hCoV-19/Denmark/DCGC-9521/2020\|EPI_ISL_669018\|2020 | ···················· | ···················· | ····················· | ···················· | ···················· | ···················· |
| T | 3154 hCoV-19/Denmark/DCGC-9523/2020\|EPI_ISL_669019\|2020 | ···················· | ···················· | ····················· | ···················· | ···················· | ···················· |
| T | 3155 hCoV-19/Denmark/DCGC-9526/2020\|EPI_ISL_669020\|2020 | ···················· | ···················· | ····················· | ···················· | ···················· | ···················· |
| T | 3156 hCoV-19/Denmark/DCGC-9529/2020\|EPI_ISL_669021\|2020 | ···················· | ···················· | ····················· | ···················· | ···················· | ···················· |
| T | 3157 hCoV-19/Denmark/DCGC-9531/2020\|EPI_ISL_669022\|2020 | ···················· | ···················· | ····················· | ···················· | ···················· | ···················· |
| T | 3158 hCoV-19/Denmark/DCGC-9532/2020\|EPI_ISL_669023\|2020 | ···················· | ···················· | ····················· | ···················· | ···················· | ···················· |
| T | 3159 hCoV-19/Denmark/DCGC-9533/2020\|EPI_ISL_669024\|2020 | ···················· | ···················· | ····················· | ···················· | ···················· | ···················· |
| T | 3160 hCoV-19/Denmark/DCGC-9536/2020\|EPI_ISL_669025\|2020 | ···················· | ·A·················· | ····················· | ···················· | ···················· | ··A················· |
| T | 3161 hCoV-19/Denmark/DCGC-9537/2020\|EPI_ISL_669026\|2020 | ···················· | ···················· | ····················· | ···················· | ···················· | ···················· |
| T | 3162 hCoV-19/Denmark/DCGC-9538/2020\|EPI_ISL_669027\|2020 | ···················· | ···················· | ····················· | ···················· | ···················· | ···················· |
| T | 3163 hCoV-19/Denmark/DCGC-9539/2020\|EPI_ISL_669028\|2020 | ···················· | ···················· | ····················· | ···················· | ···················· | ···················· |
| T | 3164 hCoV-19/Denmark/DCGC-9540/2020\|EPI_ISL_669029\|2020 | ···················· | ···················· | ····················· | ···················· | ···················· | ···················· |
| T | 3165 hCoV-19/Denmark/DCGC-9541/2020\|EPI_ISL_669030\|2020 | ···················· | ···················· | ····················· | ···················· | ···················· | ···················· |
| T | 3166 hCoV-19/Denmark/DCGC-9543/2020\|EPI_ISL_669031\|2020 | ···················· | ···················· | ····················· | ···················· | ···················· | ···················· |
| T | 3167 hCoV-19/Denmark/DCGC-9545/2020\|EPI_ISL_669032\|2020 | ···················· | ···················· | ····················· | ···················· | ···················· | ···················· |
| T | 3168 hCoV-19/Denmark/DCGC-9549/2020\|EPI_ISL_669033\|2020 | ···················· | ···················· | ····················· | ···················· | ···················· | ···················· |
| T | 3169 hCoV-19/Denmark/DCGC-9550/2020\|EPI_ISL_669034\|2020 | ···················· | ·A·················· | ····················· | ···················· | ···················· | ··A················· |
| T | 3170 hCoV-19/Denmark/DCGC-9551/2020\|EPI_ISL_669035\|2020 | ···················· | ···················· | ····················· | ···················· | ···················· | ···················· |
| T | 3171 hCoV-19/Denmark/DCGC-9554/2020\|EPI_ISL_669036\|2020 | ···················· | ···················· | ····················· | ···················· | ···················· | ···················· |
| T | 3172 hCoV-19/Denmark/DCGC-9556/2020\|EPI_ISL_669037\|2020 | ···················· | ·A·················· | ····················· | ···················· | ···················· | ···················· |
| T | 3173 hCoV-19/Denmark/DCGC-9558/2020\|EPI_ISL_669038\|2020 | ···················· | ···················· | ····················· | ···················· | ···················· | ···················· |
| T | 3174 hCoV-19/Denmark/DCGC-9559/2020\|EPI_ISL_669039\|2020 | ···················· | ···················· | ····················· | ···················· | ···················· | ···················· |
| T | 3175 hCoV-19/Denmark/DCGC-9560/2020\|EPI_ISL_669040\|2020 | ···················· | ···················· | ····················· | ···················· | ···················· | ···················· |
| T | 3176 hCoV-19/Denmark/DCGC-9563/2020\|EPI_ISL_669041\|2020 | ···················· | ···················· | ····················· | ···················· | ···················· | ···················· |
| T | 3177 hCoV-19/Denmark/DCGC-9564/2020\|EPI_ISL_669042\|2020 | ···················· | ···················· | ····················· | ···················· | ···················· | ···················· |
| T | 3178 hCoV-19/Denmark/DCGC-9565/2020\|EPI_ISL_669043\|2020 | ···················· | ···················· | ····················· | ···················· | ······T············· | ···················· |
| T | 3179 hCoV-19/Denmark/DCGC-9569/2020\|EPI_ISL_669044\|2020 | ···················· | ···················· | ····················· | ···················· | ···················· | ···················· |
| T | 3180 hCoV-19/Denmark/DCGC-9571/2020\|EPI_ISL_669045\|2020 | ···················· | ···················· | ····················· | ···················· | ···················· | ···················· |
| T | 3181 hCoV-19/Denmark/DCGC-9578/2020\|EPI_ISL_669046\|2020 | ···················· | ···················· | ····················· | ···················· | ···················· | ···················· |
| T | 3182 hCoV-19/Denmark/DCGC-9579/2020\|EPI_ISL_669047\|2020 | ···················· | ···················· | ····················· | ···················· | ···················· | ···················· |
| T | 3183 hCoV-19/Denmark/DCGC-9582/2020\|EPI_ISL_669048\|2020 | ···················· | ·A·················· | ····················· | ···················· | ···················· | ···················· |
| T | 3184 hCoV-19/Denmark/DCGC-9583/2020\|EPI_ISL_669049\|2020 | ···················· | ···················· | ····················· | ···················· | ···················· | ···················· |
| T | 3185 hCoV-19/Denmark/DCGC-9636/2020\|EPI_ISL_669080\|2020 | ···················· | ···················· | ····················· | ···················· | ···················· | ···················· |
| T | 3186 hCoV-19/Denmark/DCGC-9640/2020\|EPI_ISL_669081\|2020 | ···················· | ···················· | ····················· | ···················· | ···················· | ···················· |
| T | 3187 hCoV-19/Denmark/DCGC-9643/2020\|EPI_ISL_669082\|2020 | ···················· | ···················· | ····················· | ···················· | ···················· | ···················· |
| T | 3188 hCoV-19/Denmark/DCGC-9644/2020\|EPI_ISL_669083\|2020 | ···················· | ···················· | ····················· | ···················· | ···················· | ···················· |
| T | 3189 hCoV-19/Denmark/DCGC-9645/2020\|EPI_ISL_669084\|2020 | ···················· | ···················· | ····················· | ···················· | ···················· | ···················· |
| T | 3190 hCoV-19/Denmark/DCGC-9646/2020\|EPI_ISL_669085\|2020 | ···················· | ···················· | ····················· | ···················· | ···················· | ···················· |
| T | 3191 hCoV-19/Denmark/DCGC-9647/2020\|EPI_ISL_669086\|2020 | ···················· | ···················· | ····················· | ···················· | ···················· | ···················· |
| T | 3192 hCoV-19/Denmark/DCGC-9648/2020\|EPI_ISL_669087\|2020 | ···················· | ···················· | ····················· | ···················· | ···················· | ···················· |
| T | 3193 hCoV-19/Denmark/DCGC-9649/2020\|EPI_ISL_669088\|2020 | ···················· | ···················· | ····················· | ···················· | ···················· | ···················· |
| T | 3194 hCoV-19/Denmark/DCGC-9652/2020\|EPI_ISL_669089\|2020 | ···················· | ···················· | ····················· | ···················· | ···················· | ···················· |
| T | 3195 hCoV-19/Denmark/DCGC-9656/2020\|EPI_ISL_669090\|2020 | ···················· | ···················· | ····················· | ···················· | ···················· | ···················· |
| T | 3196 hCoV-19/Denmark/DCGC-9657/2020\|EPI_ISL_669091\|2020 | ···················· | ···················· | ····················· | ···················· | ···················· | ···················· |
| T | 3197 hCoV-19/Denmark/DCGC-9658/2020\|EPI_ISL_669092\|2020 | ···················· | ···A················ | ····················· | ···················· | ···················· | ···················· |
| T | 3198 hCoV-19/Denmark/DCGC-9661/2020\|EPI_ISL_669093\|2020 | ···················· | ···················· | ····················· | ···················· | ···················· | ···················· |

| | | | | | | | |
|---|---|---|---|---|---|---|---|
| T | 3199 hCoV-19/Denmark/DCGC-9662/2020\|EPI_ISL_669094\|2020 | ···················· | ···················· | ···················· | ···················· | ···················· | ···················· |
| T | 3200 hCoV-19/Denmark/DCGC-9663/2020\|EPI_ISL_669095\|2020 | ···················· | ···················· | ···················· | ···················· | ···················· | ···················· |
| T | 3201 hCoV-19/Denmark/DCGC-9664/2020\|EPI_ISL_669096\|2020 | ···················· | ···················· | ···················· | ···················· | ···················· | ···················· |
| T | 3202 hCoV-19/Denmark/DCGC-9665/2020\|EPI_ISL_669097\|2020 | ···················· | ···················· | ···················· | ···················· | ···················· | ···················· |
| T | 3203 hCoV-19/Denmark/DCGC-9668/2020\|EPI_ISL_669098\|2020 | ···················· | ···················· | ···················· | ···················· | ·····T·············· | ···················· |
| T | 3204 hCoV-19/Denmark/DCGC-9670/2020\|EPI_ISL_669099\|2020 | ···················· | ···················· | ···················· | ···················· | ···················· | ···················· |
| T | 3205 hCoV-19/Denmark/DCGC-10477/2020\|EPI_ISL_670000\|202 | ···················· | ···················· | ···················· | ···················· | ···················· | ···················· |
| T | 3206 hCoV-19/Denmark/DCGC-10478/2020\|EPI_ISL_670001\|202 | ···················· | ···················· | ···················· | ···················· | ···················· | ···················· |
| T | 3207 hCoV-19/Denmark/DCGC-10479/2020\|EPI_ISL_670002\|202 | ···················· | ···················· | ···················· | ···················· | ·····T·············· | ···················· |
| T | 3208 hCoV-19/Denmark/DCGC-10480/2020\|EPI_ISL_670003\|202 | ···················· | ···················· | ···················· | ···················· | ···················· | ···················· |
| T | 3209 hCoV-19/Denmark/DCGC-10481/2020\|EPI_ISL_670004\|202 | ···················· | ···················· | ···················· | ···················· | ···················· | ···················· |
| T | 3210 hCoV-19/Denmark/DCGC-10483/2020\|EPI_ISL_670005\|202 | ···················· | ·A·················· | ···················· | ···················· | ···················· | ···················· |
| T | 3211 hCoV-19/Denmark/DCGC-10484/2020\|EPI_ISL_670006\|202 | ···················· | ···················· | ···················· | ···················· | ···················· | ···················· |
| T | 3212 hCoV-19/Denmark/DCGC-10486/2020\|EPI_ISL_670007\|202 | ···················· | ···················· | ···················· | ···················· | ···················· | ···················· |
| T | 3213 hCoV-19/Denmark/DCGC-10487/2020\|EPI_ISL_670008\|202 | ···················· | ···················· | ···················· | ···················· | ···················· | ···················· |
| T | 3214 hCoV-19/Denmark/DCGC-10488/2020\|EPI_ISL_670009\|202 | ···················· | ···················· | ···················· | ···················· | ···················· | ···················· |
| T | 3215 hCoV-19/Denmark/DCGC-10489/2020\|EPI_ISL_670010\|202 | ···················· | ···················· | ···················· | ···················· | ···················· | ···················· |
| T | 3216 hCoV-19/Denmark/DCGC-10490/2020\|EPI_ISL_670011\|202 | ···················· | ···················· | ···················· | ···················· | ···················· | ···················· |
| T | 3217 hCoV-19/Denmark/DCGC-10491/2020\|EPI_ISL_670012\|202 | ···················· | ···················· | ···················· | ···················· | ···················· | ···················· |
| T | 3218 hCoV-19/Denmark/DCGC-10492/2020\|EPI_ISL_670013\|202 | ···················· | ···················· | ···················· | ···················· | ···················· | ···················· |
| T | 3219 hCoV-19/Denmark/DCGC-10493/2020\|EPI_ISL_670014\|202 | ···················· | ···················· | ···················· | ···················· | ···················· | ···················· |
| T | 3220 hCoV-19/Denmark/DCGC-10495/2020\|EPI_ISL_670015\|202 | ···················· | ···················· | ···················· | ···················· | ···················· | ···················· |
| T | 3221 hCoV-19/Denmark/DCGC-10496/2020\|EPI_ISL_670016\|202 | ···················· | ···················· | ···················· | ···················· | ···················· | ···················· |
| T | 3222 hCoV-19/Denmark/DCGC-10498/2020\|EPI_ISL_670017\|202 | ···················· | ···················· | ···················· | ···················· | ···················· | ···················· |
| T | 3223 hCoV-19/Denmark/DCGC-10499/2020\|EPI_ISL_670018\|202 | ···················· | ···················· | ···················· | ···················· | ···················· | ···················· |
| T | 3224 hCoV-19/Denmark/DCGC-10500/2020\|EPI_ISL_670019\|202 | ···················· | ···················· | ···················· | ···················· | ···················· | ···················· |
| T | 3225 hCoV-19/Denmark/DCGC-10503/2020\|EPI_ISL_670020\|202 | ···················· | ···················· | ···················· | ···················· | ···················· | ···················· |
| T | 3226 hCoV-19/Denmark/DCGC-10504/2020\|EPI_ISL_670021\|202 | ···················· | ·········A·········· | ···················· | ···················· | ···················· | ···················· |
| T | 3227 hCoV-19/Denmark/DCGC-10505/2020\|EPI_ISL_670022\|202 | ···················· | ·A·················· | ···················· | ···················· | ···················· | ···················· |
| T | 3228 hCoV-19/Denmark/DCGC-10508/2020\|EPI_ISL_670023\|202 | ···················· | ···················· | ···················· | ···················· | ···················· | ···················· |
| T | 3229 hCoV-19/Denmark/DCGC-10509/2020\|EPI_ISL_670024\|202 | ···················· | ···················· | ···················· | ···················· | ···················· | ···················· |
| T | 3230 hCoV-19/Denmark/DCGC-10510/2020\|EPI_ISL_670025\|202 | ···················· | ···················· | ···················· | ···················· | ···················· | ···················· |
| T | 3231 hCoV-19/Denmark/DCGC-10512/2020\|EPI_ISL_670026\|202 | ···················· | ···················· | ···················· | ···················· | ···················· | ···················· |
| T | 3232 hCoV-19/Denmark/DCGC-10513/2020\|EPI_ISL_670027\|202 | ···················· | ···················· | ···················· | ···················· | ···················· | ···················· |
| T | 3233 hCoV-19/Denmark/DCGC-10514/2020\|EPI_ISL_670028\|202 | ···················· | ···················· | ···················· | ···················· | ···················· | ···················· |
| T | 3234 hCoV-19/Denmark/DCGC-10515/2020\|EPI_ISL_670029\|202 | ···················· | ·A·················· | ···················· | ···················· | ···················· | ··A················· |
| T | 3235 hCoV-19/Denmark/DCGC-10517/2020\|EPI_ISL_670030\|202 | ···················· | ···················· | ···················· | ···················· | ···················· | ···················· |
| T | 3236 hCoV-19/Denmark/DCGC-10520/2020\|EPI_ISL_670031\|202 | ···················· | ···················· | ···················· | ···················· | ···················· | ···················· |
| T | 3237 hCoV-19/Denmark/DCGC-10523/2020\|EPI_ISL_670032\|202 | ···················· | ···················· | ···················· | ···················· | ···················· | ···················· |
| T | 3238 hCoV-19/Denmark/DCGC-10524/2020\|EPI_ISL_670033\|202 | ···················· | ···················· | ···················· | ···················· | ···················· | ···················· |
| T | 3239 hCoV-19/Denmark/DCGC-10580/2020\|EPI_ISL_670067\|202 | ···················· | ···················· | ···················· | ···················· | ···················· | ···················· |
| T | 3240 hCoV-19/Denmark/DCGC-10583/2020\|EPI_ISL_670068\|202 | ···················· | ···················· | ···················· | ···················· | ···················· | ···················· |
| T | 3241 hCoV-19/Denmark/DCGC-10584/2020\|EPI_ISL_670069\|202 | ···················· | ···················· | ···················· | ···················· | ···················· | ···················· |
| T | 3242 hCoV-19/Denmark/DCGC-10585/2020\|EPI_ISL_670070\|202 | ···················· | ···················· | ···················· | ···················· | ···················· | ···················· |
| T | 3243 hCoV-19/Denmark/DCGC-10588/2020\|EPI_ISL_670071\|202 | ···················· | ···················· | ···················· | ···················· | ···················· | ···················· |
| T | 3244 hCoV-19/Denmark/DCGC-10589/2020\|EPI_ISL_670072\|202 | ···················· | ···················· | ···················· | ···················· | ···················· | ···················· |
| T | 3245 hCoV-19/Denmark/DCGC-10590/2020\|EPI_ISL_670073\|202 | ···················· | ···················· | ···················· | ···················· | ···················· | ···················· |
| T | 3246 hCoV-19/Denmark/DCGC-10591/2020\|EPI_ISL_670074\|202 | ···················· | ···················· | ···················· | ···················· | ···················· | ···················· |
| T | 3247 hCoV-19/Denmark/DCGC-10594/2020\|EPI_ISL_670075\|202 | ···················· | ···················· | ···················· | ···················· | ···················· | ···················· |
| T | 3248 hCoV-19/Denmark/DCGC-10595/2020\|EPI_ISL_670076\|202 | ···················· | ·A·················· | ···················· | ···················· | ···················· | ···················· |
| T | 3249 hCoV-19/Denmark/DCGC-10596/2020\|EPI_ISL_670077\|202 | ···················· | ···················· | ···················· | ···················· | ···················· | ···················· |
| T | 3250 hCoV-19/Denmark/DCGC-10598/2020\|EPI_ISL_670078\|202 | ···················· | ···················· | ···················· | ···················· | ···················· | ···················· |
| T | 3251 hCoV-19/Denmark/DCGC-10601/2020\|EPI_ISL_670079\|202 | ···················· | ···················· | ···················· | ···················· | ···················· | ···················· |
| T | 3252 hCoV-19/Denmark/DCGC-10603/2020\|EPI_ISL_670080\|202 | ···················· | ···················· | ···················· | ···················· | ···················· | ···················· |
| T | 3253 hCoV-19/Denmark/DCGC-10604/2020\|EPI_ISL_670081\|202 | ···················· | ···················· | ···················· | ···················· | ···················· | ···················· |
| T | 3254 hCoV-19/Denmark/DCGC-10605/2020\|EPI_ISL_670082\|202 | ···················· | ···················· | ···················· | ···················· | ···················· | ···················· |
| T | 3255 hCoV-19/Denmark/DCGC-10607/2020\|EPI_ISL_670083\|202 | ···················· | ···················· | ···················· | ···················· | ···················· | ···················· |
| T | 3256 hCoV-19/Denmark/DCGC-10608/2020\|EPI_ISL_670084\|202 | ···················· | ···················· | ···················· | ···················· | ···················· | ···················· |
| T | 3257 hCoV-19/Denmark/DCGC-10609/2020\|EPI_ISL_670085\|202 | ···················· | ···················· | ···················· | ···················· | ···················· | ···················· |
| T | 3258 hCoV-19/Denmark/DCGC-10610/2020\|EPI_ISL_670086\|202 | ···················· | ···················· | ···················· | ···················· | ···················· | ···················· |
| T | 3259 hCoV-19/Denmark/DCGC-10611/2020\|EPI_ISL_670087\|202 | ···················· | ···················· | ···················· | ···················· | ···················· | ···················· |
| T | 3260 hCoV-19/Denmark/DCGC-10612/2020\|EPI_ISL_670088\|202 | ···················· | ···················· | ···················· | ···················· | ···················· | ···················· |
| T | 3261 hCoV-19/Denmark/DCGC-10613/2020\|EPI_ISL_670089\|202 | ···················· | ···················· | ···················· | ···················· | ···················· | ···················· |
| T | 3262 hCoV-19/Denmark/DCGC-10616/2020\|EPI_ISL_670090\|202 | ···················· | ·A·················· | ···················· | ···················· | ···················· | ··A················· |
| T | 3263 hCoV-19/Denmark/DCGC-10617/2020\|EPI_ISL_670091\|202 | ···················· | ···················· | ···················· | ···················· | ···················· | ···················· |
| T | 3264 hCoV-19/Denmark/DCGC-10618/2020\|EPI_ISL_670092\|202 | ···················· | ···················· | ···················· | ···················· | ···················· | ···················· |
| T | 3265 hCoV-19/Denmark/DCGC-10619/2020\|EPI_ISL_670093\|202 | ···················· | ···················· | ···················· | ···················· | ···················· | ···················· |
| T | 3266 hCoV-19/Denmark/DCGC-10620/2020\|EPI_ISL_670094\|202 | ···················· | ···················· | ···················· | ···················· | ···················· | ···················· |
| T | 3267 hCoV-19/Denmark/DCGC-10621/2020\|EPI_ISL_670095\|202 | ···················· | ·A·················· | ···················· | ···················· | ···················· | ··A················· |
| T | 3268 hCoV-19/Denmark/DCGC-10622/2020\|EPI_ISL_670096\|202 | ···················· | ···················· | ···················· | ···················· | ···················· | ···················· |
| T | 3269 hCoV-19/Denmark/DCGC-10623/2020\|EPI_ISL_670097\|202 | ···················· | ···················· | ···················· | ···················· | ···················· | ···················· |
| T | 3270 hCoV-19/Denmark/DCGC-10624/2020\|EPI_ISL_670098\|202 | ···················· | ·A·················· | ···················· | ···················· | ···················· | ···················· |
| T | 3271 hCoV-19/Denmark/DCGC-10625/2020\|EPI_ISL_670099\|202 | ···················· | ···················· | ···················· | ···················· | ···················· | ···················· |
| T | 3272 hCoV-19/Gibraltar/204841733/2020\|EPI_ISL_671300\|20 | ···················· | ···················· | ···················· | ···················· | ···················· | ···················· |
| T | 3273 hCoV-19/Gibraltar/204841734/2020\|EPI_ISL_671301\|20 | ···················· | ···················· | ···················· | ···················· | ···················· | ···················· |
| T | 3274 hCoV-19/Gibraltar/204841735/2020\|EPI_ISL_671302\|20 | ···················· | ···················· | ···················· | ···················· | ···················· | ···················· |
| T | 3275 hCoV-19/Gibraltar/204841736/2020\|EPI_ISL_671303\|20 | ···················· | ···················· | ···················· | ···················· | ···················· | ···················· |
| T | 3276 hCoV-19/Gibraltar/204841737/2020\|EPI_ISL_671304\|20 | ········Y··········· | ···················· | ···················· | ···················· | ···················· | ···················· |
| T | 3277 hCoV-19/Gibraltar/204841738/2020\|EPI_ISL_671305\|20 | ···················· | ···················· | ···················· | ···················· | ···················· | ···················· |
| T | 3278 hCoV-19/Gibraltar/204841739/2020\|EPI_ISL_671306\|20 | ···················· | ···················· | ···················· | ···················· | ···················· | ···················· |

| | | | | | | | | |
|---|---|---|---|---|---|---|---|---|
| T | 3279 hCoV-19/Gibraltar/204841741/2020\|EPI_ISL_671308\|20 | ···················· | ···················· | ···················· | ···················· | ···················· | ···················· | |
| T | 3280 hCoV-19/Gibraltar/204841742/2020\|EPI_ISL_671309\|20 | ···················· | ···················· | ···················· | ···················· | ···················· | ···················· | |
| T | 3281 hCoV-19/Gibraltar/204841743/2020\|EPI_ISL_671310\|20 | ···················· | ···················· | ···················· | ···················· | ···················· | ···················· | |
| T | 3282 hCoV-19/Gibraltar/204841745/2020\|EPI_ISL_671312\|20 | ···················· | ···················· | ···················· | ···················· | ···················· | ···················· | |
| T | 3283 hCoV-19/Gibraltar/204841746/2020\|EPI_ISL_671313\|20 | ···················· | ·A·················· | ···················· | ···················· | ···················· | ···················· | |
| T | 3284 hCoV-19/Gibraltar/204841747/2020\|EPI_ISL_671314\|20 | ···················· | ·A·················· | ···················· | ···················· | ···················· | ···················· | |
| T | 3285 hCoV-19/Gibraltar/204841748/2020\|EPI_ISL_671315\|20 | ···················· | ···················· | ···················· | ···················· | ···················· | ···················· | |
| T | 3286 hCoV-19/Gibraltar/204841749/2020\|EPI_ISL_671316\|20 | ···················· | ···················· | ···················· | ···················· | ···················· | ···················· | |
| T | 3287 hCoV-19/Gibraltar/204841750/2020\|EPI_ISL_671317\|20 | ···················· | ·A·················· | ···················· | ···················· | ············R······· | ···················· | |
| T | 3288 hCoV-19/Gibraltar/204841752/2020\|EPI_ISL_671319\|20 | ···················· | ···················· | ···················· | ···················· | ···················· | ···················· | |
| T | 3289 hCoV-19/Gibraltar/204841753/2020\|EPI_ISL_671320\|20 | ···················· | ···················· | ···················· | ···················· | ···················· | ···················· | |
| T | 3290 hCoV-19/Gibraltar/204841754/2020\|EPI_ISL_671321\|20 | ···················· | ···················· | ···················· | ···················· | ···················· | ···················· | |
| T | 3291 hCoV-19/Gibraltar/204841755/2020\|EPI_ISL_671322\|20 | ···················· | ···················· | ···················· | ···················· | ···················· | ···················· | |
| T | 3292 hCoV-19/Gibraltar/204841756/2020\|EPI_ISL_671323\|20 | ···················· | ···················· | ···················· | ···················· | ···················· | ···················· | |
| T | 3293 hCoV-19/Gibraltar/204841757/2020\|EPI_ISL_671324\|20 | ···················· | ···················· | ···················· | ···················· | ···················· | ···················· | |
| T | 3294 hCoV-19/Gibraltar/204841758/2020\|EPI_ISL_671325\|20 | ···················· | ···················· | ···················· | ···················· | ···················· | ···················· | |
| T | 3295 hCoV-19/Gibraltar/204841760/2020\|EPI_ISL_671327\|20 | ···················· | ···················· | ···················· | ···················· | ···················· | ···················· | |
| T | 3296 hCoV-19/Gibraltar/204841763/2020\|EPI_ISL_671329\|20 | ···················· | ···················· | ···················· | ···················· | ···················· | ···················· | |
| T | 3297 hCoV-19/Gibraltar/204841766/2020\|EPI_ISL_671332\|20 | ···················· | ···················· | ···················· | ···················· | ···················· | ···················· | |
| T | 3298 hCoV-19/England/204890501/2020\|EPI_ISL_671336\|2020 | ···················· | ···················· | ···················· | ···················· | ······T············· | ···················· | |
| T | 3299 hCoV-19/Ireland/D-NVRL-MPA24205/2020\|EPI_ISL_67133 | ···················· | ···················· | ···················· | ···················· | ···················· | ···················· | |
| T | 3300 hCoV-19/Ireland/D-NVRL-MPA42736/2020\|EPI_ISL_67133 | ···················· | ···················· | ···················· | ···················· | ···················· | ···················· | |
| T | 3301 hCoV-19/Ireland/D-NVRL-MPB41731/2020\|EPI_ISL_67133 | ···················· | ···················· | ···················· | ···················· | ···················· | ···················· | |
| T | 3302 hCoV-19/Ireland/D-NVRL-MPA88796/2020\|EPI_ISL_67134 | ···················· | ···················· | ···················· | ···················· | ···················· | ···················· | |
| T | 3303 hCoV-19/Ireland/D-NVRL-MPS28761/2020\|EPI_ISL_67134 | ···················· | ···················· | ···················· | ···················· | ···················· | ···················· | |
| T | 3304 hCoV-19/Ireland/WW-NVRL-MPH44355/2020\|EPI_ISL_6713 | ···················· | ···················· | ···················· | ···················· | ···················· | ···················· | |
| T | 3305 hCoV-19/Ireland/D-NVRL-MPA54621/2020\|EPI_ISL_67134 | ···················· | ···················· | ···················· | ···················· | ···················· | ···················· | |
| T | 3306 hCoV-19/Ireland/D-NVRL-MPJ65056/2020\|EPI_ISL_67134 | ···················· | ···················· | ···················· | ···················· | ···················· | ···················· | |
| T | 3307 hCoV-19/Ireland/D-NVRL-MPI39625/2020\|EPI_ISL_67134 | ···················· | ···················· | ···················· | ···················· | ···················· | ···················· | |
| T | 3308 hCoV-19/Ireland/D-NVRL-MPA31215/2020\|EPI_ISL_67134 | ···················· | ···················· | ···················· | ···················· | ···················· | ···················· | |
| T | 3309 hCoV-19/Ireland/TA-NVRL-MPO65745/2020\|EPI_ISL_6713 | ···················· | ···················· | ···················· | ···················· | ···················· | ···················· | |
| T | 3310 hCoV-19/Ireland/TA-NVRL-MPY48083/2020\|EPI_ISL_6713 | ···················· | ···················· | ···················· | ···················· | ···················· | ···················· | |
| T | 3311 hCoV-19/Ireland/WW-NVRL-MPE58928/2020\|EPI_ISL_6713 | ···················· | ····A··············· | ···················· | ···················· | ···················· | ···················· | |
| T | 3312 hCoV-19/Ireland/LK-NVRL-MPB53836/2020\|EPI_ISL_6713 | ···················· | ···················· | ···················· | ···················· | ···················· | ···················· | |
| T | 3313 hCoV-19/Ireland/LK-NVRL-MPT82886/2020\|EPI_ISL_6713 | ···················· | ···················· | ···················· | ···················· | ···················· | ···················· | |
| T | 3314 hCoV-19/Ireland/CE-NVRL-MPW96382/2020\|EPI_ISL_6713 | ···················· | ···················· | ···················· | ···················· | ···················· | ···················· | |
| T | 3315 hCoV-19/Ireland/LK-NVRL-MPH95956/2020\|EPI_ISL_6713 | ···················· | ···················· | ···················· | ···················· | ···················· | ···················· | |
| T | 3316 hCoV-19/Ireland/G-NVRL-MPJ17853/2020\|EPI_ISL_67135 | ···················· | ···················· | ···················· | ···················· | ···················· | ···················· | |
| T | 3317 hCoV-19/Ireland/D-NVRL-MPQ77261/2020\|EPI_ISL_67135 | ···················· | ···················· | ···················· | ···················· | ···················· | ···················· | |
| T | 3318 hCoV-19/Ireland/D-NVRL-MPP89189/2020\|EPI_ISL_67135 | ···················· | ···················· | ···················· | ···················· | ···················· | ···················· | |
| T | 3319 hCoV-19/Ireland/OY-NVRL-MPL81201/2020\|EPI_ISL_6713 | ···················· | ···················· | ···················· | ···················· | ···················· | ···················· | |
| T | 3320 hCoV-19/Ireland/D-NVRL-MPW83173/2020\|EPI_ISL_67135 | ···················· | ···················· | ···········T········ | ···················· | ···················· | ···················· | |
| T | 3321 hCoV-19/Ireland/D-NVRL-20Z04493/2020\|EPI_ISL_67135 | ···················· | ···················· | ···················· | ···················· | ···················· | ···················· | |
| T | 3322 hCoV-19/Ireland/D-NVRL-20Z04745/2020\|EPI_ISL_67136 | ···················· | ···················A | ···················· | ···················· | ···················· | ···················· | 1 |
| T | 3323 hCoV-19/Ireland/MN-NVRL-20Z06790/2020\|EPI_ISL_6713 | ···················· | ···················· | ···················· | ···················· | ···················· | ···················· | |
| T | 3324 hCoV-19/Ireland/D-NVRL-20Z04751/2020\|EPI_ISL_67136 | ···················· | ···················· | ···················· | ···················· | ···················· | ···················· | |
| T | 3325 hCoV-19/Ireland/D-NVRL-20Z06808/2020\|EPI_ISL_67136 | ···················· | ···················· | ···················· | ···················· | ···················· | ···················· | |
| T | 3326 hCoV-19/Ireland/D-NVRL-20Z04786/2020\|EPI_ISL_67136 | ···················· | ···················· | ···················· | ···················· | ···················· | ···················· | |
| T | 3327 hCoV-19/Ireland/D-NVRL-20Z06238/2020\|EPI_ISL_67139 | ···················· | ···················· | ···················· | ···················· | ···················· | ···················· | |
| T | 3328 hCoV-19/Ireland/D-NVRL-20Z06276/2020\|EPI_ISL_67139 | ···················· | ···················A | ···················· | ···················· | ···················· | ···················· | 1 |
| T | 3329 hCoV-19/England/QEUH-B7EBD4/2020\|EPI_ISL_661894\|20 | ···················· | ···················· | ···················· | ···················· | ···················· | ···················· | |
| T | 3330 hCoV-19/England/QEUH-B7F3CC/2020\|EPI_ISL_661899\|20 | ···················· | ···················· | ···················· | ···················· | ···················· | ···················· | |
| T | 3331 hCoV-19/England/CAMC-B787D6/2020\|EPI_ISL_661900\|20 | ···················· | ···················· | ···················· | ···················· | ···················· | ···················· | |
| T | 3332 hCoV-19/England/QEUH-B7FE52/2020\|EPI_ISL_661901\|20 | ···················· | ···················· | ···················· | ···················· | ···················· | ···················· | |
| T | 3333 hCoV-19/England/CAMC-B785CD/2020\|EPI_ISL_661903\|20 | ···················· | ···················· | ···················· | ···················· | ···················· | ···················· | |
| T | 3334 hCoV-19/England/QEUH-B7FE9E/2020\|EPI_ISL_661904\|20 | ···················· | ···················· | ···················· | ···················· | ···················· | ···················· | |
| T | 3335 hCoV-19/England/QEUH-B7EC86/2020\|EPI_ISL_661905\|20 | ···················· | ···················· | ···················· | ···················· | ···················· | ···················· | |
| T | 3336 hCoV-19/England/QEUH-B8235D/2020\|EPI_ISL_661906\|20 | ···················· | ···················· | ···················· | ···················· | ···················· | ···················· | |
| T | 3337 hCoV-19/England/QEUH-B8237B/2020\|EPI_ISL_661907\|20 | ···················· | ···················· | ···················· | ···················· | ···················· | ···················· | |
| T | 3338 hCoV-19/England/QEUH-B80C18/2020\|EPI_ISL_661908\|20 | ···················· | ···················· | ···················· | ···················· | ······T············· | ···················· | |
| T | 3339 hCoV-19/England/QEUH-B8136D/2020\|EPI_ISL_661909\|20 | ···················· | ···················· | ···················· | ···················· | ···················· | ···················· | |
| T | 3340 hCoV-19/England/QEUH-B80A3C/2020\|EPI_ISL_661910\|20 | ···················· | ···················· | ···················· | ···················· | ···················· | ···················· | |
| T | 3341 hCoV-19/England/QEUH-B812F7/2020\|EPI_ISL_661911\|20 | ···················· | ·A·················· | ···················· | ···················· | ···················· | ···················· | |
| T | 3342 hCoV-19/England/QEUH-B80823/2020\|EPI_ISL_661912\|20 | ···················· | ···················· | ···················· | ···················· | ······T············· | ···················· | |
| T | 3343 hCoV-19/England/QEUH-B7F03E/2020\|EPI_ISL_661913\|20 | ···················· | ···················· | ···················· | ···················· | ···················· | ···················· | |
| T | 3344 hCoV-19/England/QEUH-B7EFD8/2020\|EPI_ISL_661914\|20 | ···················· | ···················· | ···················· | ···················· | ···················· | ···················· | |
| T | 3345 hCoV-19/England/QEUH-B7ECD1/2020\|EPI_ISL_661916\|20 | ···················· | ···················· | ···················· | ···················· | ···················· | ···················· | |
| T | 3346 hCoV-19/England/CAMC-B77986/2020\|EPI_ISL_661917\|20 | ···················· | ···················· | ···················· | ···················· | ···················· | ···················· | |
| T | 3347 hCoV-19/England/QEUH-B828A9/2020\|EPI_ISL_661919\|20 | ···················· | ···················· | ···················· | ···················· | ···················· | ···················· | |
| T | 3348 hCoV-19/England/QEUH-B7FE16/2020\|EPI_ISL_661922\|20 | ···················· | ···················· | ···················· | ···················· | ···················· | ···················· | |
| T | 3349 hCoV-19/England/QEUH-B7FF22/2020\|EPI_ISL_661923\|20 | ···················· | ···················· | ···················· | ···················· | ······T············· | ···················· | |
| T | 3350 hCoV-19/England/QEUH-B804B6/2020\|EPI_ISL_661924\|20 | ···················· | ···················· | ···················· | ···················· | ···················· | ···················· | |
| T | 3351 hCoV-19/England/CAMC-B782A8/2020\|EPI_ISL_661925\|20 | ···················· | ·A·················· | ···················· | ···················· | ···················· | ···················· | |
| T | 3352 hCoV-19/England/CAMC-B779D1/2020\|EPI_ISL_661926\|20 | ···················· | ···················· | ···················· | ···················· | ···················· | ···················· | |
| T | 3353 hCoV-19/England/QEUH-B80762/2020\|EPI_ISL_661927\|20 | ···················· | ···················· | ···················· | ···················· | ···················· | ···················· | |
| T | 3354 hCoV-19/England/CAMC-B782C6/2020\|EPI_ISL_661928\|20 | ···················· | ···················· | ···················· | ···················· | ······T············· | ···················· | |
| T | 3355 hCoV-19/England/QEUH-B8086F/2020\|EPI_ISL_661929\|20 | ···················· | ···················· | ···················· | ···················· | ···················· | ···················· | |
| T | 3356 hCoV-19/England/CAMC-B78518/2020\|EPI_ISL_661930\|20 | ···················· | ·A·················· | ···················· | ···················· | ···················· | ···A················ | |
| T | 3357 hCoV-19/England/QEUH-B7ED29/2020\|EPI_ISL_661931\|20 | ···················· | ···················· | ···················· | ···················· | ···················· | ···················· | |
| T | 3358 hCoV-19/England/QEUH-B7EC1D/2020\|EPI_ISL_661932\|20 | ···················· | ···················· | ···················· | ···················· | ···················· | ···················· | |

T 3359 hCoV-19/England/QEUH-B80744/2020|EPI_ISL_661933|20 ···················· ···················· ···················· ···················· ···················· ····················
T 3360 hCoV-19/England/QEUH-B8287C/2020|EPI_ISL_661934|20 ···················· ···················· ···················· ···················· ···················· ····················
T 3361 hCoV-19/England/QEUH-B7FE34/2020|EPI_ISL_661935|20 ···················· ···················· ···················· ···················· ···················· ····················
T 3362 hCoV-19/England/QEUH-B7FF5F/2020|EPI_ISL_661936|20 ···················· ···················· ···················· ···················· ···················· ····················
T 3363 hCoV-19/England/QEUH-B7EF8D/2020|EPI_ISL_661937|20 ···················· ···················· ···················· ···················· ···················· ····················
T 3364 hCoV-19/England/QEUH-B80B0C/2020|EPI_ISL_661938|20 ···················· ···················· ···················· ···················· ···················· ····················
T 3365 hCoV-19/England/QEUH-B82654/2020|EPI_ISL_661939|20 ···················· ···················· ···················· ···················· ···················· ····················
T 3366 hCoV-19/England/QEUH-B82548/2020|EPI_ISL_661940|20 ···················· ···················· ···················· ···················· ···················· ····················
T 3367 hCoV-19/England/CAMC-B7883D/2020|EPI_ISL_661941|20 ···················· ···················· ···················· ···················· ······T············· ····················
T 3368 hCoV-19/England/QEUH-B7F3EA/2020|EPI_ISL_661942|20 ···················· ···················· ···················· ···················· ···················· ····················
T 3369 hCoV-19/England/CAMC-B7831E/2020|EPI_ISL_661944|20 ···················· ···················· ···················· ···················· ···················· ····················
T 3370 hCoV-19/England/QEUH-B82214/2020|EPI_ISL_661945|20 ···················· ···················· ···················· ···················· ···················· ····················
T 3371 hCoV-19/England/CAMC-B785DC/2020|EPI_ISL_661947|20 ···················· ···················· ···················· ···················· ···················· ····················
T 3372 hCoV-19/England/QEUH-B82609/2020|EPI_ISL_661948|20 ···················· ···················· ···················· ···················· ···················· ····················
T 3373 hCoV-19/England/QEUH-B8054A/2020|EPI_ISL_661949|20 ···················· ···················· ···················· ···················· ···················· ····················
T 3374 hCoV-19/England/QEUH-B82232/2020|EPI_ISL_661950|20 ···················· ···················· ···················· ···················· ···················· ····················
T 3375 hCoV-19/England/CAMC-B77BBD/2020|EPI_ISL_661951|20 ···················· ···················· ···················· ···················· ···················· ····················
T 3376 hCoV-19/England/QEUH-B80AF0/2020|EPI_ISL_661952|20 ···················· ···················· ···················· ···················· ···················· ····················
T 3377 hCoV-19/England/QEUH-B82311/2020|EPI_ISL_661955|20 ···················· ···················· ···················· ···················· ···················· ····················
T 3378 hCoV-19/England/ALDP-B7501A/2020|EPI_ISL_661956|20 ···················· ···················· ···················· ···················· ···················· ····················
T 3379 hCoV-19/England/QEUH-B68FB3/2020|EPI_ISL_661957|20 ···················· ·A·················· ···················· ···················· ···················· ··A·················
T 3380 hCoV-19/England/ALDP-B744B5/2020|EPI_ISL_661958|20 ···················· ···················· ···················· ···················· ···················· ····················
T 3381 hCoV-19/England/QEUH-B67F4B/2020|EPI_ISL_661959|20 ···················· ···················· ···················· ···················· ···················· ····················
T 3382 hCoV-19/Norway/CS-04/2020|EPI_ISL_664106|2020-04-1 ···················· ···················· ···················· ···················· ···················· ····················
T 3383 hCoV-19/England/C200042671/2020|EPI_ISL_666734|202 ···················· ·A·················· ···················· ···················· ···················· ····················
T 3384 hCoV-19/England/C200042763/2020|EPI_ISL_666735|202 ···················· ·A·················· ···················· ···················· ···················· ····················
T 3385 hCoV-19/England/204490097/2020|EPI_ISL_666740|2020 ···················· ···················· ···················· ···················· ···················· ····················
T 3386 hCoV-19/England/C200042646/2020|EPI_ISL_666741|202 ···················· ···················· ···················· ···················· ···················· ····················
T 3387 hCoV-19/England/C200043747/2020|EPI_ISL_666742|202 ···················· ···················· ···················· ···················· ···················· ····················
T 3388 hCoV-19/England/C200043779/2020|EPI_ISL_666743|202 ···················· ···················· ···················· ···················· ···················· ····················
T 3389 hCoV-19/England/C200043791/2020|EPI_ISL_666744|202 ···················· ···················· ···················· ···················· ···················· ····················
T 3390 hCoV-19/England/C200043827/2020|EPI_ISL_666745|202 ···················· ···················· ···················· ···················· ···················· ····················
T 3391 hCoV-19/England/C200043831/2020|EPI_ISL_666746|202 ···················· ···················· ···················· ···················· ···················· ····················
T 3392 hCoV-19/England/C200043921/2020|EPI_ISL_666747|202 ···················· ···················· ···················· ···················· ···················· ····················
T 3393 hCoV-19/England/C200043931/2020|EPI_ISL_666748|202 ···················· ···················· ···················· ···················· ···················· ····················
T 3394 hCoV-19/England/C200043942/2020|EPI_ISL_666749|202 ···················· ···················· ···················· ···················· ···················· ····················
T 3395 hCoV-19/England/C200043952/2020|EPI_ISL_666750|202 ···················· ···················· ···················· ···················· ···················· ····················
T 3396 hCoV-19/England/C200044010/2020|EPI_ISL_666751|202 ···················· ···················· ···················· ···················· ···················· ····················
T 3397 hCoV-19/England/C200044058/2020|EPI_ISL_666752|202 ···················· ···················· ···················· ···················· ···················· ····················
T 3398 hCoV-19/England/C200044294/2020|EPI_ISL_666753|202 ···················· ···················· ···················· ···················· ···················· ····················
T 3399 hCoV-19/England/C200044425/2020|EPI_ISL_666754|202 ···················· ···················· ···················· ···················· ···················· ····················
T 3400 hCoV-19/England/C200044442/2020|EPI_ISL_666755|202 ···················· ···················· ···················· ···················· ···················· ····················
T 3401 hCoV-19/England/C200044464/2020|EPI_ISL_666756|202 ···················· ···················· ···················· ···················· ··A················· ····················
T 3402 hCoV-19/England/C200044468/2020|EPI_ISL_666757|202 ···················· ···················· ···················· ···················· ··A················· ····················
T 3403 hCoV-19/England/C200044470/2020|EPI_ISL_666758|202 ···················· ···················· ···················· ···················· ··A················· ····················
T 3404 hCoV-19/England/C200044472/2020|EPI_ISL_666759|202 ···················· ···················· ···················· ···················· ··A················· ····················
T 3405 hCoV-19/England/C200044475/2020|EPI_ISL_666760|202 ···················· ···················· ···················· ···················· ··A················· ····················
T 3406 hCoV-19/England/C200044478/2020|EPI_ISL_666761|202 ···················· ···················· ···················· ···················· ··A················· ····················
T 3407 hCoV-19/England/C200044479/2020|EPI_ISL_666762|202 ···················· ···················· ···················· ···················· ··A················· ····················
T 3408 hCoV-19/England/C200044480/2020|EPI_ISL_666763|202 ···················· ···················· ···················· ···················· ··A················· ····················
T 3409 hCoV-19/England/C200044483/2020|EPI_ISL_666764|202 ···················· ···················· ···················· ···················· ··A················· ····················
T 3410 hCoV-19/England/C200044487/2020|EPI_ISL_666765|202 ···················· ···················· ···················· ···················· ··A················· ····················
T 3411 hCoV-19/England/C200044492/2020|EPI_ISL_666766|202 ···················· ···················· ···················· ···················· ··A················· ····················
T 3412 hCoV-19/England/C200046112/2020|EPI_ISL_666767|202 ···················· ···················· ···················· ···················· ···················· ····················
T 3413 hCoV-19/England/C200047063/2020|EPI_ISL_666768|202 ···················· ···················· ···················· ···················· ···················· ····················
T 3414 hCoV-19/England/204260135/2020|EPI_ISL_666769|2020 ···················· ···················· ···················· ···················· ···················· ····················
T 3415 hCoV-19/England/204490718/2020|EPI_ISL_666770|2020 ···················· ···················· ···················· ···················· ···················· ····················
T 3416 hCoV-19/USA/MD-MDH-0326/2020|EPI_ISL_666771|2020-1 ···················· ·A·················· ···················· ···················· ···················· ····················
T 3417 hCoV-19/USA/MD-MDH-0335/2020|EPI_ISL_666772|2020-1 ···················· ···················· ···················· ···················· ···················· ····················
T 3418 hCoV-19/USA/MD-MDH-0337/2020|EPI_ISL_666773|2020-1 ···················· ·A·················· ···················· ···················· ···················· ····················
T 3419 hCoV-19/USA/MD-MDH-0339/2020|EPI_ISL_666774|2020-1 ···················· ···················· ···················· ···················· ···················· ····················
T 3420 hCoV-19/USA/MD-MDH-0342/2020|EPI_ISL_666775|2020-1 ···················· ·A·················· ···················· ···················· ···················· ····················
T 3421 hCoV-19/USA/MD-MDH-0343/2020|EPI_ISL_666776|2020-1 ···················· ··········A········· ···················· ···················· ···················· ····················
T 3422 hCoV-19/USA/MD-MDH-0407/2020|EPI_ISL_666777|2020-1 ···················· ···················· ···················· ···················· ···················· ····················
T 3423 hCoV-19/USA/MD-MDH-0408/2020|EPI_ISL_666778|2020-1 ···················· ············C······· ···················· ···················· ···················· ····················
T 3424 hCoV-19/USA/MD-MDH-0409/2020|EPI_ISL_666779|2020-1 ···················· ············C······· ···················· ···················· ···················· ····················
T 3425 hCoV-19/USA/MD-MDH-0410/2020|EPI_ISL_666780|2020-1 ···················· ···················· ···················· ···················· ···················· ····················
T 3426 hCoV-19/USA/MD-MDH-0411/2020|EPI_ISL_666781|2020-1 ···················· ···················· ···················· ···················· ···················· ····················
T 3427 hCoV-19/USA/MD-MDH-0412/2020|EPI_ISL_666782|2020-1 ···················· ···················· ···················· ···················· ···················· ····················
T 3428 hCoV-19/USA/MD-MDH-0413/2020|EPI_ISL_666783|2020-1 ···················· ···················· ···················· ···················· ···················· ····················
T 3429 hCoV-19/USA/MD-MDH-0414/2020|EPI_ISL_666784|2020-1 ···················· ·A·················· ···················· ···················· ···················· ····················
T 3430 hCoV-19/USA/MD-MDH-0415/2020|EPI_ISL_666785|2020-1 ···················· ·············A······ ···················· ···················· ···················· ····················
T 3431 hCoV-19/USA/MD-MDH-0416/2020|EPI_ISL_666786|2020-1 ···················· ·············A······ ···················· ···················· ···················· ····················
T 3432 hCoV-19/USA/MD-MDH-0417/2020|EPI_ISL_666787|2020-1 ···················· ···················· ···········T········ ···················· ···················· ····················
T 3433 hCoV-19/USA/MD-MDH-0420/2020|EPI_ISL_666788|2020-1 ···················· ···················· ···················· ···················· ···················· ····················
T 3434 hCoV-19/USA/MD-MDH-0421/2020|EPI_ISL_666789|2020-1 ···················· ···················· ···················· ···················· ···················· ····················
T 3435 hCoV-19/USA/MD-MDH-0422/2020|EPI_ISL_666790|2020-1 ···················· ·A·················· ···················· ···················· ···················· ····················
T 3436 hCoV-19/USA/MD-MDH-0423/2020|EPI_ISL_666791|2020-1 ···················· ···················· ···················· ···················· ···················· ····················
T 3437 hCoV-19/USA/MD-MDH-0424/2020|EPI_ISL_666792|2020-1 ···················· ···················· ···················· ···················· ···················· ····················
T 3438 hCoV-19/USA/MD-MDH-0425/2020|EPI_ISL_666793|2020-1 ···················· ···················· ···················· ···················· ···················· ····················

| | | | | | | | |
|---|---|---|---|---|---|---|---|
| T | 3439 hCoV-19/USA/MD-MDH-0426/2020\|EPI_ISL_666794\|2020-1 | ···················· | ·A·················· | ···················· | ···················· | ···················· | ···················· |
| T | 3440 hCoV-19/USA/MD-MDH-0427/2020\|EPI_ISL_666795\|2020-1 | ···················· | ·A·················· | ···················· | ···················· | ···················· | ···················· |
| T | 3441 hCoV-19/USA/MD-MDH-0428/2020\|EPI_ISL_666796\|2020-1 | ···················· | ·A·················· | ···················· | ···················· | ···················· | ···················· |
| T | 3442 hCoV-19/USA/MD-MDH-0429/2020\|EPI_ISL_666797\|2020-1 | ···················· | ·A·················· | ···················· | ···················· | ···················· | ···················· |
| T | 3443 hCoV-19/USA/MD-MDH-0430/2020\|EPI_ISL_666798\|2020-1 | ···················· | ···················· | ···················· | ···················· | ···················· | ···················· |
| T | 3444 hCoV-19/USA/MD-MDH-0431/2020\|EPI_ISL_666799\|2020-1 | ···················· | ···················· | ···················· | ···················· | ···················· | ···················· |
| T | 3445 hCoV-19/USA/MD-MDH-0443/2020\|EPI_ISL_666810\|2020-1 | ···················· | ···················· | ···················· | ···················· | ···················· | ···················· |
| T | 3446 hCoV-19/Denmark/DCGC-9584/2020\|EPI_ISL_669050\|2020 | ···················· | ···················· | ···················· | ···················· | ···················· | ···················· |
| T | 3447 hCoV-19/Denmark/DCGC-9587/2020\|EPI_ISL_669051\|2020 | ···················· | ···················· | ···················· | ···················· | ···················· | ···················· |
| T | 3448 hCoV-19/Denmark/DCGC-9591/2020\|EPI_ISL_669052\|2020 | ···················· | ···················· | ···················· | ···················· | ···················· | ···················· |
| T | 3449 hCoV-19/Denmark/DCGC-9593/2020\|EPI_ISL_669053\|2020 | ···················· | ···················· | ···················· | ···················· | ···················· | ···················· |
| T | 3450 hCoV-19/Denmark/DCGC-9594/2020\|EPI_ISL_669054\|2020 | ···················· | ···················· | ···················· | ···················· | ···················· | ···················· |
| T | 3451 hCoV-19/Denmark/DCGC-9595/2020\|EPI_ISL_669055\|2020 | ···················· | ···················· | ···················· | ···················· | ···················· | ···················· |
| T | 3452 hCoV-19/Denmark/DCGC-9596/2020\|EPI_ISL_669056\|2020 | ···················· | ···················· | ···················· | ···················· | ···················· | ···················· |
| T | 3453 hCoV-19/Denmark/DCGC-9597/2020\|EPI_ISL_669057\|2020 | ···················· | ···················· | ···················· | ···················· | ···················· | ···················· |
| T | 3454 hCoV-19/Denmark/DCGC-9599/2020\|EPI_ISL_669058\|2020 | ···················· | ···················· | ···················· | ···················· | ···················· | ···················· |
| T | 3455 hCoV-19/Denmark/DCGC-9600/2020\|EPI_ISL_669059\|2020 | ···················· | ···················· | ···················· | ···················· | ···················· | ···················· |
| T | 3456 hCoV-19/Denmark/DCGC-9602/2020\|EPI_ISL_669060\|2020 | ···················· | ···················· | ···················· | ···················· | ···················· | ···················· |
| T | 3457 hCoV-19/Denmark/DCGC-9603/2020\|EPI_ISL_669061\|2020 | ···················· | ···················· | ···················· | ···················· | ···················· | ···················· |
| T | 3458 hCoV-19/Denmark/DCGC-9604/2020\|EPI_ISL_669062\|2020 | ···················· | ···················· | ···················· | ···················· | ···················· | ···················· |
| T | 3459 hCoV-19/Denmark/DCGC-9605/2020\|EPI_ISL_669063\|2020 | ···················· | ·A·················· | ···················· | ···················· | ···················· | ···A················ |
| T | 3460 hCoV-19/Denmark/DCGC-9606/2020\|EPI_ISL_669064\|2020 | ···················· | ···················· | ···················· | ···················· | ···················· | ···················· |
| T | 3461 hCoV-19/Denmark/DCGC-9610/2020\|EPI_ISL_669065\|2020 | ···················· | ···················· | ···················· | ···················· | ·······T············ | ···················· |
| T | 3462 hCoV-19/Denmark/DCGC-9612/2020\|EPI_ISL_669066\|2020 | ···················· | ···················· | ···················· | ···················· | ···················· | ···················· |
| T | 3463 hCoV-19/Denmark/DCGC-9613/2020\|EPI_ISL_669067\|2020 | ···················· | ···················· | ···················· | ···················· | ···················· | ···················· |
| T | 3464 hCoV-19/Denmark/DCGC-9614/2020\|EPI_ISL_669068\|2020 | ···················· | ···················· | ···················· | ···················· | ···················· | ···················· |
| T | 3465 hCoV-19/Denmark/DCGC-9616/2020\|EPI_ISL_669069\|2020 | ···················· | ···················· | ···················· | ···················· | ···················· | ···················· |
| T | 3466 hCoV-19/Denmark/DCGC-9619/2020\|EPI_ISL_669070\|2020 | ···················· | ···················· | ···················· | ···················· | ···················· | ···················· |
| T | 3467 hCoV-19/Denmark/DCGC-9621/2020\|EPI_ISL_669071\|2020 | ···················· | ···················· | ···················· | ···················· | ···················· | ···················· |
| T | 3468 hCoV-19/Denmark/DCGC-9625/2020\|EPI_ISL_669072\|2020 | ···················· | ···················· | ···················· | ···················· | ···················· | ···················· |
| T | 3469 hCoV-19/Denmark/DCGC-9626/2020\|EPI_ISL_669073\|2020 | ···················· | ···················· | ···················· | ···················· | ···················· | ···················· |
| T | 3470 hCoV-19/Denmark/DCGC-9627/2020\|EPI_ISL_669074\|2020 | ···················· | ···················· | ···················· | ···················· | ···················· | ···················· |
| T | 3471 hCoV-19/Denmark/DCGC-9628/2020\|EPI_ISL_669075\|2020 | ···················· | ···················· | ···················· | ···················· | ···················· | ···················· |
| T | 3472 hCoV-19/Denmark/DCGC-9629/2020\|EPI_ISL_669076\|2020 | ···················· | ···················· | ···················· | ···················· | ···················· | ···················· |
| T | 3473 hCoV-19/Denmark/DCGC-9632/2020\|EPI_ISL_669077\|2020 | ···················· | ···················· | ···················· | ···················· | ···················· | ···················· |
| T | 3474 hCoV-19/Denmark/DCGC-9634/2020\|EPI_ISL_669078\|2020 | ···················· | ···················· | ···················· | ···················· | ···················· | ···················· |
| T | 3475 hCoV-19/Denmark/DCGC-9635/2020\|EPI_ISL_669079\|2020 | ···················· | ···················· | ···················· | ···················· | ···················· | ···················· |
| T | 3476 hCoV-19/Denmark/DCGC-10526/2020\|EPI_ISL_670034\|202 | ···················· | ···················· | ···················· | ···················· | ···················· | ···················· |
| T | 3477 hCoV-19/Denmark/DCGC-10528/2020\|EPI_ISL_670035\|202 | ···················· | ···················· | ···················· | ···················· | ···················· | ···················· |
| T | 3478 hCoV-19/Denmark/DCGC-10530/2020\|EPI_ISL_670036\|202 | ···················· | ···················· | ···················· | ···················· | ···················· | ···················· |
| T | 3479 hCoV-19/Denmark/DCGC-10531/2020\|EPI_ISL_670037\|202 | ···················· | ···················· | ···················· | ···················· | ···················· | ···················· |
| T | 3480 hCoV-19/Denmark/DCGC-10538/2020\|EPI_ISL_670038\|202 | ···················· | ···················· | ···················· | ···················· | ···················· | ···················· |
| T | 3481 hCoV-19/Denmark/DCGC-10540/2020\|EPI_ISL_670039\|202 | ···················· | ···················· | ···················· | ···················· | ···················· | ···················· |
| T | 3482 hCoV-19/Denmark/DCGC-10541/2020\|EPI_ISL_670040\|202 | ···················· | ···················· | ···················· | ···················· | ···················· | ···················· |
| T | 3483 hCoV-19/Denmark/DCGC-10542/2020\|EPI_ISL_670041\|202 | ···················· | ···················· | ···················· | ···················· | ···················· | ···················· |
| T | 3484 hCoV-19/Denmark/DCGC-10543/2020\|EPI_ISL_670042\|202 | ···················· | ···················· | ···················· | ···················· | ···················· | ···················· |
| T | 3485 hCoV-19/Denmark/DCGC-10546/2020\|EPI_ISL_670043\|202 | ···················· | ···················· | ···················· | ···················· | ···················· | ···················· |
| T | 3486 hCoV-19/Denmark/DCGC-10547/2020\|EPI_ISL_670044\|202 | ···················· | ···················· | ···················· | ···················· | ···················· | ···················· |
| T | 3487 hCoV-19/Denmark/DCGC-10548/2020\|EPI_ISL_670045\|202 | ···················· | ···················· | ···················· | ···················· | ···················· | ···················· |
| T | 3488 hCoV-19/Denmark/DCGC-10549/2020\|EPI_ISL_670046\|202 | ···················· | ···················· | ···················· | ···················· | ···················· | ···················· |
| T | 3489 hCoV-19/Denmark/DCGC-10550/2020\|EPI_ISL_670047\|202 | ···················· | ···················· | ···················· | ···················· | ···················· | ···················· |
| T | 3490 hCoV-19/Denmark/DCGC-10551/2020\|EPI_ISL_670048\|202 | ···················· | ···················· | ···················· | ···················· | ···················· | ···················· |
| T | 3491 hCoV-19/Denmark/DCGC-10553/2020\|EPI_ISL_670049\|202 | ···················· | ···················· | ···················· | ···················· | ···················· | ···················· |
| T | 3492 hCoV-19/Denmark/DCGC-10555/2020\|EPI_ISL_670050\|202 | ···················· | ···················· | ···················· | ···················· | ···················· | ···················· |
| T | 3493 hCoV-19/Denmark/DCGC-10557/2020\|EPI_ISL_670051\|202 | ···················· | ···················· | ···················· | ···················· | ···················· | ···················· |
| T | 3494 hCoV-19/Denmark/DCGC-10559/2020\|EPI_ISL_670052\|202 | ···················· | ···················· | ···················· | ···················· | ···················· | ···················· |
| T | 3495 hCoV-19/Denmark/DCGC-10563/2020\|EPI_ISL_670053\|202 | ···················· | ···················· | ···················· | ···················· | ···················· | ···················· |
| T | 3496 hCoV-19/Denmark/DCGC-10566/2020\|EPI_ISL_670054\|202 | ···················· | ·A·················· | ···················· | ···················· | ···················· | ···················· |
| T | 3497 hCoV-19/Denmark/DCGC-10568/2020\|EPI_ISL_670055\|202 | ···················· | ···················· | ···················· | ···················· | ···················· | ···················· |
| T | 3498 hCoV-19/Denmark/DCGC-10569/2020\|EPI_ISL_670056\|202 | ···················· | ···················· | ···················· | ···················· | ···················· | ···················· |
| T | 3499 hCoV-19/Denmark/DCGC-10570/2020\|EPI_ISL_670057\|202 | ···················· | ···················· | ···················· | ···················· | ···················· | ···················· |
| T | 3500 hCoV-19/Denmark/DCGC-10571/2020\|EPI_ISL_670058\|202 | ···················· | ···················· | ···················· | ···················· | ···················· | ···················· |
| T | 3501 hCoV-19/Denmark/DCGC-10572/2020\|EPI_ISL_670059\|202 | ···················· | ···················· | ···················· | ···················· | ···················· | ···················· |
| T | 3502 hCoV-19/Denmark/DCGC-10573/2020\|EPI_ISL_670060\|202 | ···················· | ·A·················· | ···················· | ···················· | ···················· | ···················· |
| T | 3503 hCoV-19/Denmark/DCGC-10574/2020\|EPI_ISL_670061\|202 | ···················· | ···················· | ···················· | ···················· | ···················· | ···················· |
| T | 3504 hCoV-19/Denmark/DCGC-10575/2020\|EPI_ISL_670062\|202 | ···················· | ···················· | ···················· | ···················· | ···················· | ···················· |
| T | 3505 hCoV-19/Denmark/DCGC-10576/2020\|EPI_ISL_670063\|202 | ···················· | ···················· | ···················· | ···················· | ···················· | ···················· |
| T | 3506 hCoV-19/Denmark/DCGC-10577/2020\|EPI_ISL_670064\|202 | ···················· | ···················· | ···················· | ···················· | ···················· | ···················· |
| T | 3507 hCoV-19/Denmark/DCGC-10578/2020\|EPI_ISL_670065\|202 | ···················· | ···················· | ···················· | ···················· | ···················· | ···················· |
| T | 3508 hCoV-19/Denmark/DCGC-10579/2020\|EPI_ISL_670066\|202 | ···················· | ·A·················· | ···················· | ···················· | ···················· | ···················· |
| T | 3509 hCoV-19/Denmark/DCGC-11923/2020\|EPI_ISL_671219\|202 | ···················· | ···················· | ···················· | ···················· | ···················· | ···················· |
| T | 3510 hCoV-19/Denmark/DCGC-11930/2020\|EPI_ISL_671222\|202 | ···················· | ···················· | ·········T·········· | ···················· | ···················· | ···················· |
| T | 3511 hCoV-19/Denmark/DCGC-11931/2020\|EPI_ISL_671223\|202 | ···················· | ···················· | ···················· | ···················· | ···················· | ···················· |
| T | 3512 hCoV-19/Denmark/DCGC-11932/2020\|EPI_ISL_671224\|202 | ···················· | ···················· | ···················· | ···················· | ···················· | ···················· |
| T | 3513 hCoV-19/Denmark/DCGC-11935/2020\|EPI_ISL_671225\|202 | ···················· | ···················· | ···················· | ···················· | ···················· | ···················· |
| T | 3514 hCoV-19/Denmark/DCGC-11940/2020\|EPI_ISL_671226\|202 | ···················· | ···················· | ···················· | ···················· | ···················· | ···················· |
| T | 3515 hCoV-19/Denmark/DCGC-11943/2020\|EPI_ISL_671227\|202 | ···················· | ···················· | ···················· | ···················· | ···················· | ···················· |
| T | 3516 hCoV-19/Denmark/DCGC-11946/2020\|EPI_ISL_671228\|202 | ···················· | ···················· | ···················· | ···················· | ···················· | ···················· |
| T | 3517 hCoV-19/Denmark/DCGC-11948/2020\|EPI_ISL_671229\|202 | ···················· | ···················· | ···················· | ···················· | ···················· | ···················· |
| T | 3518 hCoV-19/Denmark/DCGC-11956/2020\|EPI_ISL_671230\|202 | ···················· | ···················· | ···················· | ···················· | ···················· | ···················· |

| | | | | | | | | |
|---|---|---|---|---|---|---|---|---|
| T | 3519 hCoV-19/Denmark/DCGC-11958/2020\|EPI_ISL_671231\|202 | ···················· | ···················· | ···················· | ···················· | ···················· | ···················· | |
| T | 3520 hCoV-19/Denmark/DCGC-11959/2020\|EPI_ISL_671232\|202 | ···················· | ···················· | ···················· | ···················· | ···················· | ···················· | |
| T | 3521 hCoV-19/Denmark/DCGC-11965/2020\|EPI_ISL_671233\|202 | ···················· | ···················· | ···················· | ···················· | ···················· | ···················· | |
| T | 3522 hCoV-19/Denmark/DCGC-11976/2020\|EPI_ISL_671234\|202 | ········T··········· | ···················· | ···················· | ···················· | ···················· | ···················· | |
| T | 3523 hCoV-19/Denmark/DCGC-11981/2020\|EPI_ISL_671235\|202 | ···················· | ···················· | ···················· | ···················· | ···················· | ···················· | |
| T | 3524 hCoV-19/Denmark/DCGC-11984/2020\|EPI_ISL_671236\|202 | ···················· | ···················· | ···················· | ···················· | ···················· | ···················· | |
| T | 3525 hCoV-19/Denmark/DCGC-11988/2020\|EPI_ISL_671237\|202 | ········T··········· | ···················· | ···················· | ···················· | ···················· | ···················· | |
| T | 3526 hCoV-19/Denmark/DCGC-11989/2020\|EPI_ISL_671238\|202 | ········T··········· | ···················· | ···················· | ···················· | ···················· | ···················· | |
| T | 3527 hCoV-19/Denmark/DCGC-11990/2020\|EPI_ISL_671239\|202 | ········T··········· | ···················· | ···················· | ···················· | ···················· | ···················· | |
| T | 3528 hCoV-19/Denmark/DCGC-11992/2020\|EPI_ISL_671240\|202 | ········T··········· | ···················· | ···················· | ···················· | ···················· | ···················· | |
| T | 3529 hCoV-19/Denmark/DCGC-11994/2020\|EPI_ISL_671241\|202 | ···················· | ···················· | ···················· | ···················· | ···················· | ···················· | |
| T | 3530 hCoV-19/Denmark/DCGC-11995/2020\|EPI_ISL_671242\|202 | ···················· | ·A·················· | ···················· | ···················· | ···················· | ···A················ | |
| T | 3531 hCoV-19/Denmark/DCGC-11996/2020\|EPI_ISL_671243\|202 | ···················· | ···················· | ···················· | ···················· | ···················· | ···················· | |
| T | 3532 hCoV-19/Ireland/LH-NVRL-20Z06931/2020\|EPI_ISL_6713 | ···················· | ···················· | ···················· | ···················· | ···················· | ···················· | |
| T | 3533 hCoV-19/Ireland/MO-NVRL-20Z04470/2020\|EPI_ISL_6713 | ···················· | ···················· | ···················· | ···················· | ···················· | ···················· | |
| T | 3534 hCoV-19/Ireland/MN-NVRL-20Z06661/2020\|EPI_ISL_6713 | ···················· | ···················· | ···················· | ···················· | ···················· | ···················· | |
| T | 3535 hCoV-19/Ireland/MN-NVRL-20Z06657/2020\|EPI_ISL_6713 | ···················· | ···················· | ···················· | ···················· | ···················· | ···················· | |
| T | 3536 hCoV-19/Ireland/MO-NVRL-20Z04451/2020\|EPI_ISL_6713 | ···················· | ···················· | ···················· | ···················· | ···················· | ···················· | |
| T | 3537 hCoV-19/Ireland/D-NVRL-20Z04363/2020\|EPI_ISL_67137 | ···················· | ···················· | ···················· | ···················· | ···················· | ···················· | |
| T | 3538 hCoV-19/Ireland/WH-NVRL-20Z06323/2020\|EPI_ISL_6713 | ···················· | ···················· | ···················· | ···················· | ···················· | ···················· | |
| T | 3539 hCoV-19/Ireland/WW-NVRL-20Z06465/2020\|EPI_ISL_6713 | ···················· | ···················· | ···················· | ···················· | ···················· | ···················· | |
| T | 3540 hCoV-19/Ireland/D-NVRL-20G41470/2020\|EPI_ISL_67137 | ···················· | ···················· | ···················· | ···················· | ···················· | ···················· | |
| T | 3541 hCoV-19/Ireland/D-NVRL-20G41469/2020\|EPI_ISL_67137 | ···················· | ···················· | ···················· | ···················· | ···················· | ···················· | |
| T | 3542 hCoV-19/Ireland/KE-NVRL-20G27987/2020\|EPI_ISL_6713 | ···················· | ···················· | ···················· | ···················· | ···················· | ···················· | |
| T | 3543 hCoV-19/Ireland/D-NVRL-20G38988/2020\|EPI_ISL_67137 | ···················· | ···················A | ···················· | ···················· | ···················· | ···················· | 1 |
| T | 3544 hCoV-19/Ireland/MH-NVRL-20G44544/2020\|EPI_ISL_6713 | ···················· | ···················· | ···················· | ···················· | ···················· | ···················· | |
| T | 3545 hCoV-19/Ireland/MH-NVRL-20G44537/2020\|EPI_ISL_6713 | ···················· | ···················A | ···················· | ···················· | ···················· | ···················· | 1 |
| T | 3546 hCoV-19/Ireland/MN-NVRL-20G40668/2020\|EPI_ISL_6713 | ···················· | ···················· | ···················· | ···················· | ···················· | ···················· | |
| T | 3547 hCoV-19/Ireland/MN-NVRL-20G40665/2020\|EPI_ISL_6713 | ···················· | ···················· | ···················· | ···················· | ···················· | ···················· | |
| T | 3548 hCoV-19/Ireland/LS-NVRL-20Z06636/2020\|EPI_ISL_6713 | ···················· | ···················· | ···················· | ···················· | ···················· | ···················· | |
| T | 3549 hCoV-19/Ireland/D-NVRL-20Z06343/2020\|EPI_ISL_67138 | ···················· | ···················· | ···················· | ···················· | ···················· | ···················· | |
| T | 3550 hCoV-19/Ireland/D-NVRL-20Z04313/2020\|EPI_ISL_67138 | ···················· | ···················· | ···················· | ···················· | ···················· | ···················· | |
| T | 3551 hCoV-19/Ireland/D-NVRL-20Z06353/2020\|EPI_ISL_67138 | ···················· | ···················· | ···················· | ···················· | ···················· | ···················· | |
| T | 3552 hCoV-19/Ireland/LS-NVRL-20Z06376/2020\|EPI_ISL_6713 | ···················· | ···················· | ···················· | ···················· | ···················· | ···················· | |
| T | 3553 hCoV-19/Ireland/D-NVRL-20Z04315/2020\|EPI_ISL_67138 | ···················· | ···················· | ···················· | ···················· | ···················· | ···················· | |
| T | 3554 hCoV-19/Ireland/D-NVRL-20Z06397/2020\|EPI_ISL_67138 | ···················· | ···················· | ···················· | ···················· | ···················· | ···················· | |
| T | 3555 hCoV-19/Ireland/D-NVRL-20Z06270/2020\|EPI_ISL_67138 | ···················· | ···················A | ···················· | ···················· | ···················· | ···················· | 1 |
| T | 3556 hCoV-19/Ireland/WW-NVRL-20Z06456/2020\|EPI_ISL_6713 | ···················· | ···················· | ···················· | ···················· | ···················· | ···················· | |
| T | 3557 hCoV-19/Ireland/D-NVRL-20Z06275/2020\|EPI_ISL_67139 | ···················· | ···················A | ···················· | ···················· | ···················· | ···················· | 1 |
| T | 3558 hCoV-19/Ireland/MN-NVRL-20Z06686/2020\|EPI_ISL_6713 | ···················· | ···················· | ···················· | ···················· | ···················· | ···················· | |
| T | 3559 hCoV-19/Ireland/WW-NVRL-20Z06457/2020\|EPI_ISL_6713 | ···················· | ···················· | ···················· | ···················· | ···················· | ···················· | |
| T | 3560 hCoV-19/Ireland/WW-NVRL-20Z06466/2020\|EPI_ISL_6713 | ···················· | ···················· | ···················· | ···················· | ···················· | ···················· | |
| T | 3561 hCoV-19/Ireland/MN-NVRL-20Z06210/2020\|EPI_ISL_6713 | ···················· | ···················· | ···················· | ···················· | ···················· | ···················· | |
| T | 3562 hCoV-19/Ireland/D-NVRL-20Z04316/2020\|EPI_ISL_67139 | ···················· | ···················· | ···················· | ···················· | ···················· | ···················· | |
| T | 3563 hCoV-19/Ireland/MN-NVRL-20Z06212/2020\|EPI_ISL_6713 | ···················· | ·············A······ | ···················· | ···················· | ···················· | ···················· | |
| T | 3564 hCoV-19/Ireland/D-NVRL-20Z06274/2020\|EPI_ISL_67139 | ···················· | ···················A | ···················· | ···················· | ···················· | ···················· | 1 |
| T | 3565 hCoV-19/USA/UT-UPHL-201203314/2020\|EPI_ISL_672553\| | ···················· | ···················· | ···················· | ···················· | ···················· | ···················· | |
| T | 3566 hCoV-19/USA/UT-UPHL-201206253/2020\|EPI_ISL_672554\| | ···················· | ···················· | ···················· | ···················· | ···················· | ···················· | |
| T | 3567 hCoV-19/USA/UT-UPHL-201219634/2020\|EPI_ISL_672555\| | ···················· | ···················· | ···················· | ···················· | ···················· | ···················· | |
| T | 3568 hCoV-19/USA/UT-UPHL-201203016/2020\|EPI_ISL_672556\| | ···················· | ···················· | ···················· | ···················· | ···················· | ···················· | |
| T | 3569 hCoV-19/USA/UT-UPHL-201232095/2020\|EPI_ISL_672557\| | ···················· | ···················· | ···················· | ···················· | ···················· | ···················· | |
| T | 3570 hCoV-19/USA/UT-UPHL-201228458/2020\|EPI_ISL_672558\| | ···················· | ···················· | ···················· | ···················· | ···················· | ···················· | |
| T | 3571 hCoV-19/USA/UT-UPHL-201212641/2020\|EPI_ISL_672559\| | ···················· | ···················· | ···················· | ···················· | ···················· | ···················· | |
| T | 3572 hCoV-19/USA/UT-UPHL-201205227/2020\|EPI_ISL_672560\| | ···················· | ···················· | ···················· | ···················· | ···················· | ···················· | |
| T | 3573 hCoV-19/USA/UT-UPHL-201208296/2020\|EPI_ISL_672561\| | ···················· | ···················· | ···················· | ···················· | ···················· | ···················· | |
| T | 3574 hCoV-19/USA/UT-UPHL-201227098/2020\|EPI_ISL_672562\| | ···················· | ···················· | ···················· | ···················· | ···················· | ···················· | |
| T | 3575 hCoV-19/USA/UT-UPHL-201200759/2020\|EPI_ISL_672563\| | ···················· | ···················· | ···················· | ···················· | ···················· | ···················· | |
| T | 3576 hCoV-19/USA/UT-UPHL-201207011/2020\|EPI_ISL_672564\| | ···················· | ···················· | ···················· | ···················· | ······T············· | ···················· | |
| T | 3577 hCoV-19/USA/UT-UPHL-201226976/2020\|EPI_ISL_672565\| | ···················· | ···················· | ···················· | ···················· | ···················· | ···················· | |
| T | 3578 hCoV-19/USA/UT-UPHL-201228183/2020\|EPI_ISL_672566\| | ···················· | ···················· | ···················· | ···················· | ···················· | ···················· | |
| T | 3579 hCoV-19/USA/UT-UPHL-201220800/2020\|EPI_ISL_672567\| | ···················· | ···················· | ···················· | ···················· | ···················· | ···················· | |
| T | 3580 hCoV-19/USA/UT-UPHL-201230409/2020\|EPI_ISL_672568\| | ···················· | ···················· | ···················· | ···················· | ···················· | ···················· | |
| T | 3581 hCoV-19/USA/UT-UPHL-201224344/2020\|EPI_ISL_672569\| | ···················· | ···················· | ···················· | ···················· | ···················· | ···················· | |
| T | 3582 hCoV-19/El Salvador/UES-04/2020\|EPI_ISL_672570\|202 | ···················· | ···················· | ···················· | ···················· | ···················· | ···················· | |
| T | 3583 hCoV-19/El Salvador/UES-05/2020\|EPI_ISL_672572\|202 | ···················· | ···················· | ···················· | ···················· | ···················· | ···················· | |
| T | 3584 hCoV-19/El Salvador/UES-INS-06/2020\|EPI_ISL_672573 | ···················· | ···················· | ···················· | ···················· | ···················· | ···················· | |
| T | 3585 hCoV-19/England/QEUH-B8102A/2020\|EPI_ISL_661840\|20 | ···················· | ···················· | ···················· | ···················· | ···················· | ···················· | |
| T | 3586 hCoV-19/England/QEUH-B7FDFB/2020\|EPI_ISL_661841\|20 | ···················· | ···················· | ···················· | ···················· | ···················· | ···················· | |
| T | 3587 hCoV-19/England/QEUH-B8289A/2020\|EPI_ISL_661842\|20 | ···················· | ···················· | ···················· | ···················· | ···················· | ···················· | |
| T | 3588 hCoV-19/England/QEUH-B827D9/2020\|EPI_ISL_661846\|20 | ···················· | ···················· | ···················· | ···················· | ···················· | ···················· | |
| T | 3589 hCoV-19/England/QEUH-B80A78/2020\|EPI_ISL_661847\|20 | ···················· | ···················· | ···················· | ···················· | ···················· | ···················· | |
| T | 3590 hCoV-19/England/QEUH-B805B3/2020\|EPI_ISL_661848\|20 | ···················· | ···················· | ···················· | ···················· | ···················· | ···················· | |
| T | 3591 hCoV-19/England/QEUH-B813A9/2020\|EPI_ISL_661849\|20 | ···················· | ···················· | ···················· | ···················· | ···················· | ···················· | |
| T | 3592 hCoV-19/England/QEUH-B826FA/2020\|EPI_ISL_661850\|20 | ···················· | ···················· | ···················· | ···················· | ···················· | ···················· | |
| T | 3593 hCoV-19/England/QEUH-B809D5/2020\|EPI_ISL_661851\|20 | ···················· | ···················· | ···················· | ···················· | ···················· | ···················· | |
| T | 3594 hCoV-19/England/QEUH-B813B8/2020\|EPI_ISL_661852\|20 | ···················· | ···················· | ···················· | ···················· | ···················· | ···················· | |
| T | 3595 hCoV-19/England/CAMC-B77B62/2020\|EPI_ISL_661853\|20 | ···················· | ···················· | ···················· | ···················· | ···················· | ···················· | |
| T | 3596 hCoV-19/England/QEUH-B822BA/2020\|EPI_ISL_661855\|20 | ···················· | ···················· | ···················· | ···················· | ···················· | ···················· | |
| T | 3597 hCoV-19/England/CAMC-B77B80/2020\|EPI_ISL_661856\|20 | ···················· | ···················· | ···················· | ···················· | ···················· | ···················· | |
| T | 3598 hCoV-19/England/QEUH-B7F04D/2020\|EPI_ISL_661857\|20 | ···················· | ···················· | ···················· | ···················· | ···················· | ···················· | |

| | | | | | | | | |
|---|---|---|---|---|---|---|---|---|
| T | 3599 | hCoV-19/England/QEUH-B7FF31/2020\|EPI_ISL_661859\|20 | .................... | .................... | .................... | .................... | .................... | .................... |
| T | 3600 | hCoV-19/England/QEUH-B827AC/2020\|EPI_ISL_661860\|20 | .................... | .................... | .................... | .................... | .................... | .................... |
| T | 3601 | hCoV-19/England/QEUH-B822AB/2020\|EPI_ISL_661861\|20 | .................... | .................... | .................... | .................... | .................... | .................... |
| T | 3602 | hCoV-19/England/QEUH-B80568/2020\|EPI_ISL_661862\|20 | .................... | .A.................. | .................... | .................... | .................... | .................... |
| T | 3603 | hCoV-19/England/CAMC-B784DF/2020\|EPI_ISL_661863\|20 | .................... | .................... | .................... | .................... | .................... | .................... |
| T | 3604 | hCoV-19/England/QEUH-B8284F/2020\|EPI_ISL_661865\|20 | .................... | .................... | .................... | .................... | .................... | .................... |
| T | 3605 | hCoV-19/England/QEUH-B7EC77/2020\|EPI_ISL_661866\|20 | .................... | .................... | .................... | .................... | .................... | .................... |
| T | 3606 | hCoV-19/England/QEUH-B80FD3/2020\|EPI_ISL_661867\|20 | .................... | .................... | .................... | .................... | .................... | .................... |
| T | 3607 | hCoV-19/England/QEUH-B7FEE9/2020\|EPI_ISL_661868\|20 | .................... | .................... | .................... | .................... | .................... | .................... |
| T | 3608 | hCoV-19/England/CAMC-B7792C/2020\|EPI_ISL_661869\|20 | .................... | .................... | .................... | .................... | ......T............. | .................... |
| T | 3609 | hCoV-19/England/QEUH-B80586/2020\|EPI_ISL_661870\|20 | .................... | .................... | .................... | .................... | .................... | .................... |
| T | 3610 | hCoV-19/England/QEUH-B825B1/2020\|EPI_ISL_661872\|20 | .................... | .................... | .................... | .................... | .................... | .................... |
| T | 3611 | hCoV-19/England/CAMC-B77B71/2020\|EPI_ISL_661873\|20 | .................... | .................... | .................... | .................... | ....T............... | .................... |
| T | 3612 | hCoV-19/England/QEUH-B8050E/2020\|EPI_ISL_661875\|20 | .................... | .................... | .................... | .................... | .................... | .................... |
| T | 3613 | hCoV-19/England/QEUH-B82681/2020\|EPI_ISL_661877\|20 | .................... | .................... | .................... | .................... | .................... | .................... |
| T | 3614 | hCoV-19/England/QEUH-B7F06B/2020\|EPI_ISL_661878\|20 | .................... | .................... | .................... | .................... | .................... | .................... |
| T | 3615 | hCoV-19/England/QEUH-B807DB/2020\|EPI_ISL_661879\|20 | .................... | .................... | .................... | .................... | .................... | .................... |
| T | 3616 | hCoV-19/England/QEUH-B8233F/2020\|EPI_ISL_661880\|20 | .................... | .................... | .................... | .................... | .................... | .................... |
| T | 3617 | hCoV-19/England/CAMC-B77537/2020\|EPI_ISL_661881\|20 | .................... | .................... | .................... | .................... | .................... | .................... |
| T | 3618 | hCoV-19/England/QEUH-B82803/2020\|EPI_ISL_661884\|20 | .................... | .................... | .................... | .................... | .................... | .................... |
| T | 3619 | hCoV-19/England/QEUH-B7F05C/2020\|EPI_ISL_661885\|20 | .................... | .................... | .................... | .................... | .................... | .................... |
| T | 3620 | hCoV-19/England/CAMC-B78554/2020\|EPI_ISL_661887\|20 | .................... | .................... | .................... | .................... | .................... | .................... |
| T | 3621 | hCoV-19/England/QEUH-B825EE/2020\|EPI_ISL_661888\|20 | .................... | .................... | .................... | .................... | .................... | .................... |
| T | 3622 | hCoV-19/England/QEUH-B7FE25/2020\|EPI_ISL_661889\|20 | .................... | .................... | .................... | .................... | .................... | .................... |
| T | 3623 | hCoV-19/England/QEUH-B7EFE7/2020\|EPI_ISL_661890\|20 | .................... | .................... | .................... | .................... | .................... | .................... |
| T | 3624 | hCoV-19/England/QEUH-B8279D/2020\|EPI_ISL_661891\|20 | .................... | .................... | .................... | .................... | .................... | .................... |
| T | 3625 | hCoV-19/England/QEUH-B813D6/2020\|EPI_ISL_661893\|20 | .................... | .................... | .................... | .................... | .................... | .................... |
| T | 3626 | hCoV-19/Australia/VIC17695/2020\|EPI_ISL_664000\|202 | .................... | .................... | .................... | .................... | .................... | .................... |
| T | 3627 | hCoV-19/Australia/VIC17696/2020\|EPI_ISL_664001\|202 | .................... | .................... | .................... | .................... | .................... | .................... |
| T | 3628 | hCoV-19/Australia/VIC17702/2020\|EPI_ISL_664002\|202 | .................... | .................... | .................... | .................... | .................... | .................... |
| T | 3629 | hCoV-19/Australia/VIC17705/2020\|EPI_ISL_664003\|202 | .................... | .................... | .................... | .................... | .................... | .................... |
| T | 3630 | hCoV-19/Australia/VIC17710/2020\|EPI_ISL_664004\|202 | .................... | .................... | .................... | .................... | .................... | .................... |
| T | 3631 | hCoV-19/Australia/VIC17711/2020\|EPI_ISL_664005\|202 | .................... | .................... | .................... | .................... | .................... | .................... |
| T | 3632 | hCoV-19/Australia/VIC17712/2020\|EPI_ISL_664006\|202 | .................... | .................... | .................... | .................... | .................... | .................... |
| T | 3633 | hCoV-19/Australia/VIC17713/2020\|EPI_ISL_664007\|202 | .................... | .................... | .................... | .................... | .................... | .................... |
| T | 3638 | hCoV-19/Australia/VIC17723/2020\|EPI_ISL_664012\|202 | .................... | .................... | .................... | .................... | .................... | .................... |
| T | 3639 | hCoV-19/Australia/VIC17724/2020\|EPI_ISL_664013\|202 | .................... | .................... | .................... | .................... | .................... | .................... |
| T | 3643 | hCoV-19/Australia/VIC17728/2020\|EPI_ISL_664017\|202 | .................... | .................... | .................... | .................... | .................... | .................... |
| T | 3644 | hCoV-19/Australia/VIC17732/2020\|EPI_ISL_664018\|202 | .................... | .................... | .................... | .................... | .................... | .................... |
| T | 3645 | hCoV-19/Australia/VIC17733/2020\|EPI_ISL_664019\|202 | .................... | .................... | .................... | .................... | .................... | .................... |
| T | 3646 | hCoV-19/Germany/NW-HHU-322/2020\|EPI_ISL_666624\|202 | .................... | .................... | .................... | .................... | .................... | .................... |
| T | 3647 | hCoV-19/Germany/NW-HHU-325/2020\|EPI_ISL_666626\|202 | .................... | .................... | .................... | .................... | .................... | .................... |
| T | 3648 | hCoV-19/Germany/NW-HHU-326/2020\|EPI_ISL_666627\|202 | .................... | .................... | .................... | .................... | .................... | .................... |
| T | 3649 | hCoV-19/Germany/NW-HHU-350/2020\|EPI_ISL_666637\|202 | .................... | .................... | .................... | .................... | .................... | .................... |
| T | 3650 | hCoV-19/Germany/NW-HHU-353/2020\|EPI_ISL_666640\|202 | .........T.......... | .................... | .................... | .................... | .................... | .................... |
| T | 3651 | hCoV-19/Germany/NW-HHU-355/2020\|EPI_ISL_666642\|202 | .................... | .................... | .................... | .................... | .................... | .................... |
| T | 3652 | hCoV-19/Germany/NW-HHU-361/2020\|EPI_ISL_666648\|202 | .................... | .................... | .................... | .................... | .................... | .................... |
| T | 3653 | hCoV-19/France/ARA-SC019/2020\|EPI_ISL_666654\|2020- | .................... | .................... | .................... | .................... | .................... | .................... |
| T | 3654 | hCoV-19/France/ARA-SC026/2020\|EPI_ISL_666656\|2020- | .................... | .................... | .................... | .................... | .................... | .................... |
| T | 3655 | hCoV-19/France/ARA-SC029/2020\|EPI_ISL_666657\|2020- | .................... | .................... | .................... | .................... | .................... | .................... |
| T | 3656 | hCoV-19/France/ARA-SC031/2020\|EPI_ISL_666659\|2020- | .................... | .................... | .................... | .................... | .................... | .................... |
| T | 3657 | hCoV-19/France/ARA-SC037/2020\|EPI_ISL_666660\|2020- | .................... | .................... | .................... | .................... | .................... | .................... |
| T | 3658 | hCoV-19/France/ARA-SC038/2020\|EPI_ISL_666661\|2020- | .................... | .................... | .................... | .................... | .................... | .................... |
| T | 3659 | hCoV-19/France/ARA-SC041/2020\|EPI_ISL_666662\|2020- | .................... | .................... | .................... | .................... | .................... | .................... |
| T | 3660 | hCoV-19/France/ARA-SC044/2020\|EPI_ISL_666664\|2020- | .................... | .................... | .................... | .................... | .................... | .................... |
| T | 3661 | hCoV-19/France/ARA-SC045/2020\|EPI_ISL_666665\|2020- | .................... | .................... | .................... | .................... | .................... | .................... |
| T | 3662 | hCoV-19/France/ARA-SC049/2020\|EPI_ISL_666668\|2020- | .................... | .................... | .................... | .................... | .................... | .................... |
| T | 3663 | hCoV-19/France/ARA-SC055/2020\|EPI_ISL_666669\|2020- | .................... | .................... | .................... | .................... | .................... | .................... |
| T | 3665 | hCoV-19/France/ARA-SC057/2020\|EPI_ISL_666671\|2020- | .................... | .................... | .................... | .................... | .................... | .................... |
| T | 3666 | hCoV-19/France/ARA-SC059/2020\|EPI_ISL_666672\|2020- | .................... | .................... | .................... | .................... | .................... | .................... |
| T | 3667 | hCoV-19/France/ARA-SC061/2020\|EPI_ISL_666673\|2020- | .................... | .................... | .................... | .................... | .................... | .................... |
| T | 3669 | hCoV-19/France/ARA-SC067/2020\|EPI_ISL_666676\|2020- | .................... | .................... | .................... | .................... | .................... | .................... |
| T | 3670 | hCoV-19/France/ARA-SC069/2020\|EPI_ISL_666677\|2020- | .................... | .................... | .................... | .................... | .................... | .................... |
| T | 3671 | hCoV-19/France/ARA-SC071/2020\|EPI_ISL_666678\|2020- | .................... | .................... | .................... | .................... | .................... | .................... |
| T | 3672 | hCoV-19/France/ARA-SC074/2020\|EPI_ISL_666679\|2020- | .................... | .................... | .................... | .................... | .................... | .................... |
| T | 3673 | hCoV-19/France/ARA-SC077/2020\|EPI_ISL_666680\|2020- | .................... | .................... | .................... | .................... | .................... | .................... |
| T | 3674 | hCoV-19/France/ARA-SC080/2020\|EPI_ISL_666681\|2020- | .................... | .................... | .................... | .................... | .................... | .................... |
| T | 3675 | hCoV-19/France/ARA-SC097/2020\|EPI_ISL_666683\|2020- | .................... | .................... | .................... | .................... | .................... | .................... |
| T | 3676 | hCoV-19/France/ARA-SC098/2020\|EPI_ISL_666684\|2020- | .................... | .................... | .................... | .................... | .................... | .................... |
| T | 3677 | hCoV-19/France/ARA-SC099/2020\|EPI_ISL_666685\|2020- | .................... | .................... | .................... | .................... | .................... | .................... |
| T | 3678 | hCoV-19/France/ARA-SC106/2020\|EPI_ISL_666686\|2020- | .................... | .............A...... | .................... | .................... | .................... | .................... |
| T | 3679 | hCoV-19/Ireland/DL-NVRL-20G49337/2020\|EPI_ISL_6666 | .................... | .................... | .................... | .................... | .................... | .................... |
| T | 3680 | hCoV-19/Ireland/DL-NVRL-20G49554/2020\|EPI_ISL_6666 | .................... | .................... | .................... | .................... | .................... | .................... |
| T | 3681 | hCoV-19/Ireland/DL-NVRL-20G49141/2020\|EPI_ISL_6666 | .................... | .................... | .................... | .................... | .................... | .................... |
| T | 3682 | hCoV-19/Ireland/DL-NVRL-20G48972/2020\|EPI_ISL_6666 | .................... | .................... | .................... | .................... | .................... | .................... |
| T | 3683 | hCoV-19/Ireland/DL-NVRL-20G48992/2020\|EPI_ISL_6667 | .................... | .................... | .................... | .................... | .................... | .................... |
| T | 3684 | hCoV-19/Ireland/DL-NVRL-20G48985/2020\|EPI_ISL_6667 | .................... | .................... | .................... | .................... | .................... | .................... |
| T | 3685 | hCoV-19/Ireland/DL-NVRL-20G48996/2020\|EPI_ISL_6667 | .................... | .................... | .................... | .................... | .................... | .................... |
| T | 3686 | hCoV-19/Ireland/DL-NVRL-20G45634/2020\|EPI_ISL_6667 | .................... | .................... | .................... | .................... | .................... | .................... |
| T | 3687 | hCoV-19/Ireland/DL-NVRL-20G48997/2020\|EPI_ISL_6667 | .................... | .................... | .................... | .................... | .................... | .................... |

| | | | | | | | | |
|---|---|---|---|---|---|---|---|---|
| T | 3688 hCoV-19/Ireland/DL-NVRL-20G45114/2020\|EPI_ISL_6667 | ···················· | ···················· | ····················· | ···················· | ···················· | ···················· | |
| T | 3689 hCoV-19/Ireland/DL-NVRL-20G45120/2020\|EPI_ISL_6667 | ···················· | ···················· | ····················· | ···················· | ···················· | ···················· | |
| T | 3690 hCoV-19/Ireland/DL-NVRL-20G45117/2020\|EPI_ISL_6667 | ···················· | ···················· | ····················· | ···················· | ···················· | ···················· | |
| T | 3691 hCoV-19/Ireland/DL-NVRL-20G45123/2020\|EPI_ISL_6667 | ···················· | ···················· | ····················· | ···················· | ···················· | ···················· | |
| T | 3692 hCoV-19/Ireland/KE-NVRL-20G49144/2020\|EPI_ISL_6667 | ···················· | ···················· | ····················· | ···················· | ···················· | ···················· | |
| T | 3693 hCoV-19/Ireland/MO-NVRL-20G48980/2020\|EPI_ISL_6667 | ···················· | ···················· | ····················· | ···················· | ···················· | ···················· | |
| T | 3694 hCoV-19/France/NAQ-SC290/2020\|EPI_ISL_666711\|2020- | ···················· | ···················· | ····················· | ···················· | ···················· | ···················· | |
| T | 3695 hCoV-19/France/NAQ-SC291/2020\|EPI_ISL_666712\|2020- | ···················· | ···················A | ····················· | ···················· | ···················· | ···················· | 1 |
| T | 3696 hCoV-19/France/NAQ-SC292/2020\|EPI_ISL_666713\|2020- | ···················· | ···················· | ····················· | ···················· | ···················· | ···················· | |
| T | 3697 hCoV-19/France/NAQ-SC293/2020\|EPI_ISL_666714\|2020- | ···················· | ···················· | ····················· | ···················· | ···················· | ···················· | |
| T | 3698 hCoV-19/France/NAQ-SC294/2020\|EPI_ISL_666715\|2020- | ···················· | ···················· | ····················· | ···················· | ···················· | ···················· | |
| T | 3700 hCoV-19/France/NAQ-SC296/2020\|EPI_ISL_666717\|2020- | ···················· | ···················· | ····················· | ···················· | ···················· | ···················· | |
| T | 3701 hCoV-19/France/NAQ-SC298/2020\|EPI_ISL_666718\|2020- | ···················· | ···················· | ····················· | ···················· | ···················· | ···················· | |
| T | 3702 hCoV-19/France/NAQ-SC300/2020\|EPI_ISL_666719\|2020- | ···················· | ···················· | ····················· | ···················· | ···················· | ···················· | |
| T | 3703 hCoV-19/France/NAQ-SC301/2020\|EPI_ISL_666720\|2020- | ···················· | ···················· | ····················· | ···················· | ···················· | ···················· | |
| T | 3704 hCoV-19/France/PDL-SC652/2020\|EPI_ISL_666721\|2020- | ···················· | ···················· | ····················· | ···················· | ···················· | ···················· | |
| T | 3705 hCoV-19/France/PDL-SC653/2020\|EPI_ISL_666722\|2020- | ···················· | ···················· | ····················· | ···················· | ···················· | ···················· | |
| T | 3706 hCoV-19/France/PDL-SC656/2020\|EPI_ISL_666723\|2020- | ···················· | ···················· | ····················· | ···················· | ···················· | ···················· | |
| T | 3707 hCoV-19/France/PDL-SC658/2020\|EPI_ISL_666724\|2020- | ···················· | ···················· | ····················· | ···················· | ···················· | ···················· | |
| T | 3708 hCoV-19/England/C200042605/2020\|EPI_ISL_666731\|202 | ···················· | ·A·················· | ····················· | ···················· | ···················· | ···················· | |
| T | 3709 hCoV-19/England/C200042606/2020\|EPI_ISL_666732\|202 | ···················· | ·A·················· | ····················· | ···················· | ···················· | ···················· | |
| T | 3710 hCoV-19/England/C200042607/2020\|EPI_ISL_666733\|202 | ···················· | ·A·················· | ····················· | ···················· | ···················· | ···················· | |
| T | 3711 hCoV-19/England/CAMC-B797E4/2020\|EPI_ISL_667955\|20 | ···················· | ···················· | ····················· | ···················· | ···················· | ···················· | |
| T | 3712 hCoV-19/England/QEUH-B7E59A/2020\|EPI_ISL_667958\|20 | ···················· | ···················· | ····················· | ···················· | ···················· | ···················· | |
| T | 3713 hCoV-19/England/QEUH-B7E9E9/2020\|EPI_ISL_667966\|20 | ···················· | ···················· | ····················· | ···················· | ···················· | ···················· | |
| T | 3714 hCoV-19/England/QEUH-B80F79/2020\|EPI_ISL_667967\|20 | ···················· | ···················· | ····················· | ···················· | ···················· | ···················· | |
| T | 3715 hCoV-19/England/CAMC-B7967E/2020\|EPI_ISL_667968\|20 | ···················· | ···················· | ····················· | ···················· | ···················· | ···················· | |
| T | 3716 hCoV-19/England/QEUH-B7EE80/2020\|EPI_ISL_667969\|20 | ···················· | ···················· | ····················· | ···················· | ···················· | ···················· | |
| T | 3717 hCoV-19/England/QEUH-B81AA4/2020\|EPI_ISL_667970\|20 | ···················· | ···················· | ····················· | ···················· | ···················· | ···················· | |
| T | 3718 hCoV-19/England/CAMC-B79984/2020\|EPI_ISL_667971\|20 | ···················· | ···················· | ····················· | ···················· | ···················· | ···················· | |
| T | 3719 hCoV-19/England/QEUH-B7E58B/2020\|EPI_ISL_667972\|20 | ···················· | ···················· | ····················· | ···················· | ···················· | ···················· | |
| T | 3720 hCoV-19/England/QEUH-B7EA9B/2020\|EPI_ISL_667973\|20 | ···················· | ···················· | ····················· | ···················· | ···················· | ···················· | |
| T | 3721 hCoV-19/England/QEUH-B7EE71/2020\|EPI_ISL_667975\|20 | ···················· | ···················· | ····················· | ···················· | ···················· | ···················· | |
| T | 3722 hCoV-19/England/QEUH-B7EEEA/2020\|EPI_ISL_667977\|20 | ···················· | ···················· | ····················· | ···················· | ···················· | ···················· | |
| T | 3723 hCoV-19/England/QEUH-B7ED74/2020\|EPI_ISL_667978\|20 | ···················· | ···················· | ····················· | ···················· | ···················· | ···················· | |
| T | 3724 hCoV-19/England/CAMC-B79A09/2020\|EPI_ISL_667979\|20 | ···················· | ···················· | ····················· | ···················· | ···················· | ···················· | |
| T | 3725 hCoV-19/England/QEUH-B7ED83/2020\|EPI_ISL_667983\|20 | ···················· | ···················· | ····················· | ···················· | ···················· | ···················· | |
| T | 3726 hCoV-19/England/CAMC-B79B9D/2020\|EPI_ISL_667985\|20 | ···················· | ···················· | ····················· | ···················· | ······T············· | ···················· | |
| T | 3727 hCoV-19/England/QEUH-B7E5D6/2020\|EPI_ISL_667986\|20 | ···················· | ···················· | ····················· | ···················· | ···················· | ···················· | |
| T | 3728 hCoV-19/England/QEUH-B81725/2020\|EPI_ISL_667989\|20 | ···················· | ···················· | ····················· | ···················· | ···················· | ···················· | |
| T | 3729 hCoV-19/England/QEUH-B81B1A/2020\|EPI_ISL_667990\|20 | ···················· | ···················· | ····················· | ···················· | ···················· | ···················· | |
| T | 3730 hCoV-19/England/CAMC-B78C5F/2020\|EPI_ISL_667991\|20 | ···················· | ···················· | ····················· | ···················· | ···················· | ···················· | |
| T | 3731 hCoV-19/England/QEUH-B81628/2020\|EPI_ISL_667992\|20 | ···················· | ···················· | ····················· | ···················· | ···················· | ···················· | |
| T | 3732 hCoV-19/England/CAMC-B78A19/2020\|EPI_ISL_667993\|20 | ···················· | ···················· | ····················· | ···················· | ···················· | ···A················ | |
| T | 3733 hCoV-19/England/CAMC-B79BCA/2020\|EPI_ISL_667994\|20 | ···················· | ···················· | ····················· | ···················· | ···················· | ···················· | |
| T | 3734 hCoV-19/England/CAMC-B79E0D/2020\|EPI_ISL_667995\|20 | ···················· | ···················· | ····················· | ···················· | ···················· | ···················· | |
| T | 3735 hCoV-19/England/QEUH-B80E8B/2020\|EPI_ISL_667996\|20 | ···················· | ···················· | ····················· | ···················· | ···················· | ···················· | |
| T | 3736 hCoV-19/Denmark/DCGC-11891/2020\|EPI_ISL_671200\|202 | ···················· | ···················· | ····················· | ···················· | ···················· | ···················· | |
| T | 3737 hCoV-19/Denmark/DCGC-11894/2020\|EPI_ISL_671201\|202 | ···················· | ·A·················· | ····················· | ···················· | ···················· | ···················· | |
| T | 3738 hCoV-19/Denmark/DCGC-11895/2020\|EPI_ISL_671202\|202 | ···················· | ···················· | ····················· | ···················· | ···················· | ···················· | |
| T | 3739 hCoV-19/Denmark/DCGC-11896/2020\|EPI_ISL_671203\|202 | ···················· | ···················· | ····················· | ···················· | ···················· | ···················· | |
| T | 3740 hCoV-19/Denmark/DCGC-11898/2020\|EPI_ISL_671204\|202 | ···················· | ···················· | ····················· | ···················· | ···················· | ···················· | |
| T | 3741 hCoV-19/Denmark/DCGC-11900/2020\|EPI_ISL_671205\|202 | ···················· | ···················· | ····················· | ···················· | ···················· | ···················· | |
| T | 3742 hCoV-19/Denmark/DCGC-11901/2020\|EPI_ISL_671206\|202 | ···················· | ···················· | ····················· | ···················· | ···················· | ···················· | |
| T | 3743 hCoV-19/Denmark/DCGC-11902/2020\|EPI_ISL_671207\|202 | ···················· | ···················· | ····················· | ···················· | ···················· | ···················· | |
| T | 3744 hCoV-19/Denmark/DCGC-11903/2020\|EPI_ISL_671208\|202 | ···················· | ···················· | ····················· | ···················· | ···················· | ···················· | |
| T | 3745 hCoV-19/Denmark/DCGC-11904/2020\|EPI_ISL_671209\|202 | ···················· | ···················· | ····················· | ···················· | ···················· | ···················· | |
| T | 3746 hCoV-19/Denmark/DCGC-11905/2020\|EPI_ISL_671210\|202 | ···················· | ·A·················· | ····················· | ···················· | ···················· | ···················· | |
| T | 3747 hCoV-19/Denmark/DCGC-11907/2020\|EPI_ISL_671211\|202 | ···················· | ···················· | ····················· | ···················· | ···················· | ···················· | |
| T | 3748 hCoV-19/Denmark/DCGC-11909/2020\|EPI_ISL_671212\|202 | ···················· | ···················· | ····················· | ···················· | ···················· | ···················· | |
| T | 3749 hCoV-19/Denmark/DCGC-11910/2020\|EPI_ISL_671213\|202 | ···················· | ···················· | ····················· | ···················· | ···················· | ···················· | |
| T | 3750 hCoV-19/Denmark/DCGC-11911/2020\|EPI_ISL_671214\|202 | ···················· | ···················· | ····················· | ···················· | ···················· | ···················· | |
| T | 3751 hCoV-19/Denmark/DCGC-11913/2020\|EPI_ISL_671215\|202 | ···················· | ···················· | ····················· | ···················· | ···················· | ···················· | |
| T | 3752 hCoV-19/Denmark/DCGC-11916/2020\|EPI_ISL_671216\|202 | ···················· | ···················· | ····················· | ···················· | ···················· | ···················· | |
| T | 3753 hCoV-19/Denmark/DCGC-11917/2020\|EPI_ISL_671217\|202 | ···················· | ···················· | ····················· | ···················· | ···················· | ···················· | |
| T | 3754 hCoV-19/Denmark/DCGC-11918/2020\|EPI_ISL_671218\|202 | ···················· | ·A·················· | ····················· | ···················· | ···················· | ···················· | |
| T | 3755 hCoV-19/Denmark/DCGC-11927/2020\|EPI_ISL_671220\|202 | ···················· | ···················· | ····················· | ···················· | ···················· | ···················· | |
| T | 3756 hCoV-19/Denmark/DCGC-11929/2020\|EPI_ISL_671221\|202 | ···················· | ···················· | ····················· | ···················· | ···················· | ···················· | |
| T | 3757 hCoV-19/Denmark/DCGC-11997/2020\|EPI_ISL_671244\|202 | ···················· | ···················· | ····················· | ···················· | ···················· | ···················· | |
| T | 3758 hCoV-19/Denmark/DCGC-11998/2020\|EPI_ISL_671245\|202 | ···················· | ···················· | ····················· | ···················· | ···················· | ···················· | |
| T | 3759 hCoV-19/Denmark/DCGC-11999/2020\|EPI_ISL_671246\|202 | ···················· | ···················· | ····················· | ···················· | ···················· | ···················· | |
| T | 3760 hCoV-19/Denmark/DCGC-12000/2020\|EPI_ISL_671247\|202 | ···················· | ···················· | ····················· | ···················· | ···················· | ···················· | |
| T | 3761 hCoV-19/Denmark/DCGC-12001/2020\|EPI_ISL_671248\|202 | ···················· | ···················· | ····················· | ···················· | ···················· | ···················· | |
| T | 3762 hCoV-19/Denmark/DCGC-12002/2020\|EPI_ISL_671249\|202 | ···················· | ···················· | ····················· | ···················· | ···················· | ···················· | |
| T | 3763 hCoV-19/Denmark/DCGC-12003/2020\|EPI_ISL_671250\|202 | ···················· | ···················· | ····················· | ···················· | ···················· | ···················· | |
| T | 3764 hCoV-19/Denmark/DCGC-12004/2020\|EPI_ISL_671251\|202 | ···················· | ···················· | ····················· | ···················· | ···················· | ···················· | |
| T | 3765 hCoV-19/Denmark/DCGC-12005/2020\|EPI_ISL_671252\|202 | ···················· | ···················· | ····················· | ···················· | ···················· | ···················· | |
| T | 3766 hCoV-19/Denmark/DCGC-12006/2020\|EPI_ISL_671253\|202 | ···················· | ···················· | ····················· | ···················· | ···················· | ···················· | |
| T | 3767 hCoV-19/Denmark/DCGC-12007/2020\|EPI_ISL_671254\|202 | ···················· | ···················· | ····················· | ···················· | ···················· | ···················· | |
| T | 3768 hCoV-19/Denmark/DCGC-12008/2020\|EPI_ISL_671255\|202 | ···················· | ···················· | ····················· | ···················· | ···················· | ···················· | |

| | | | | | | | |
|---|---|---|---|---|---|---|---|
| T | 3769 hCoV-19/Denmark/DCGC-12009/2020\|EPI_ISL_671256\|202 | ···················· | ···················· | ···················· | ···················· | ···················· | ···················· |
| T | 3770 hCoV-19/Denmark/DCGC-12010/2020\|EPI_ISL_671257\|202 | ···················· | ···················· | ···················· | ···················· | ···················· | ···················· |
| T | 3771 hCoV-19/Denmark/DCGC-12011/2020\|EPI_ISL_671258\|202 | ···················· | ···················· | ···················· | ···················· | ···················· | ···················· |
| T | 3772 hCoV-19/Denmark/DCGC-12012/2020\|EPI_ISL_671259\|202 | ···················· | ···················· | ···················· | ···················· | ···················· | ···················· |
| T | 3773 hCoV-19/Denmark/DCGC-12013/2020\|EPI_ISL_671260\|202 | ···················· | ···················· | ···················· | ···················· | ···················· | ···················· |
| T | 3774 hCoV-19/Denmark/DCGC-12014/2020\|EPI_ISL_671261\|202 | ···················· | ···················· | ···················· | ···················· | ···················· | ···················· |
| T | 3775 hCoV-19/Denmark/DCGC-12015/2020\|EPI_ISL_671262\|202 | ···················· | ···················· | ···················· | ···················· | ···················· | ···················· |
| T | 3776 hCoV-19/Denmark/DCGC-12016/2020\|EPI_ISL_671263\|202 | ···················· | ···················· | ···················· | ···················· | ···················· | ···················· |
| T | 3777 hCoV-19/Denmark/DCGC-12017/2020\|EPI_ISL_671264\|202 | ···················· | ···················· | ···················· | ···················· | ···················· | ···················· |
| T | 3778 hCoV-19/Denmark/DCGC-12018/2020\|EPI_ISL_671265\|202 | ···················· | ···················· | ···················· | ···················· | ···················· | ···················· |
| T | 3779 hCoV-19/Denmark/DCGC-12019/2020\|EPI_ISL_671266\|202 | ···················· | ···················· | ···················· | ···················· | ···················· | ···················· |
| T | 3780 hCoV-19/Denmark/DCGC-12020/2020\|EPI_ISL_671267\|202 | ···················· | ···················· | ···················· | ···················· | ···················· | ···················· |
| T | 3781 hCoV-19/Denmark/DCGC-12021/2020\|EPI_ISL_671268\|202 | ···················· | ···················· | ···················· | ···················· | ···················· | ···················· |
| T | 3782 hCoV-19/Denmark/DCGC-12023/2020\|EPI_ISL_671269\|202 | ···················· | ···················· | ···················· | ···················· | ···················· | ···················· |
| T | 3783 hCoV-19/Gibraltar/204841699/2020\|EPI_ISL_671270\|20 | ···················· | ···················· | ···················· | ···················· | ···················· | ···················· |
| T | 3784 hCoV-19/Gibraltar/204841700/2020\|EPI_ISL_671271\|20 | ···················· | ···················· | ···················· | ···················· | ···················· | ···················· |
| T | 3785 hCoV-19/Gibraltar/204841702/2020\|EPI_ISL_671273\|20 | ···················· | ···················· | ···················· | ···················· | ···················· | ···················· |
| T | 3786 hCoV-19/Gibraltar/204841703/2020\|EPI_ISL_671274\|20 | ···················· | ···················· | ···················· | ···················· | ···················· | ···················· |
| T | 3787 hCoV-19/Gibraltar/204841704/2020\|EPI_ISL_671275\|20 | ···················· | ···················· | ···················· | ···················· | ···················· | ···················· |
| T | 3788 hCoV-19/Gibraltar/204841705/2020\|EPI_ISL_671276\|20 | ···················· | ·A·················· | ···················· | ···················· | ···················· | ···················· |
| T | 3789 hCoV-19/Gibraltar/204841706/2020\|EPI_ISL_671277\|20 | ···················· | ···················· | ···················· | ···················· | ···················· | ···················· |
| T | 3790 hCoV-19/Gibraltar/204841707/2020\|EPI_ISL_671278\|20 | ···················· | ···················· | ···················· | ···················· | ···················· | ···················· |
| T | 3791 hCoV-19/Gibraltar/204841709/2020\|EPI_ISL_671280\|20 | ···················· | ·A·················· | ···················· | ···················· | ···················· | ···················· |
| T | 3792 hCoV-19/Gibraltar/204841710/2020\|EPI_ISL_671281\|20 | ···················· | ···················· | ···················· | ···················· | ···················· | ···················· |
| T | 3793 hCoV-19/Gibraltar/204841711/2020\|EPI_ISL_671282\|20 | ···················· | ···················· | ···················· | ···················· | ···················· | ···················· |
| T | 3794 hCoV-19/Gibraltar/204841714/2020\|EPI_ISL_671285\|20 | ···················· | ···················· | ···················· | ···················· | ···················· | ···················· |
| T | 3795 hCoV-19/Gibraltar/204841717/2020\|EPI_ISL_671286\|20 | ···················· | ···················· | ···················· | ···················· | ···················· | ···················· |
| T | 3796 hCoV-19/Gibraltar/204841718/2020\|EPI_ISL_671287\|20 | ···················· | ···················· | ···················· | ···················· | ···················· | ···················· |
| T | 3797 hCoV-19/Gibraltar/204841720/2020\|EPI_ISL_671289\|20 | ···················· | ···················· | ···················· | ···················· | ···················· | ···················· |
| T | 3798 hCoV-19/Gibraltar/204841721/2020\|EPI_ISL_671290\|20 | ···················· | ·A·················· | ···················· | ···················· | ···················· | ···················· |
| T | 3799 hCoV-19/Gibraltar/204841722/2020\|EPI_ISL_671291\|20 | ···················· | ·A·················· | ···················· | ···················· | ···················· | ···················· |
| T | 3800 hCoV-19/Gibraltar/204841723/2020\|EPI_ISL_671292\|20 | ···················· | ···················· | ···················· | ···················· | ···················· | ···················· |
| T | 3801 hCoV-19/Gibraltar/204841725/2020\|EPI_ISL_671293\|20 | ···················· | ·A·················· | ···················· | ···················· | ···················· | ···················· |
| T | 3802 hCoV-19/Gibraltar/204841726/2020\|EPI_ISL_671294\|20 | ···················· | ·A·················· | ···················· | ···················· | ···················· | ···················· |
| T | 3803 hCoV-19/Gibraltar/204841728/2020\|EPI_ISL_671296\|20 | ···················· | ···················· | ···················· | ···················· | ···················· | ···················· |
| T | 3804 hCoV-19/Gibraltar/204841730/2020\|EPI_ISL_671297\|20 | ···················· | ···················· | ···················· | ···················· | ···················· | ···················· |
| T | 3805 hCoV-19/Gibraltar/204841732/2020\|EPI_ISL_671299\|20 | ···················· | ···················· | ···················· | ···················· | ···················· | ···················· |
| T | 3807 hCoV-19/USA/CA-CZB-13468/2020\|EPI_ISL_672501\|2020- | ···················· | ···················· | ···················· | ···················· | ···················· | ···················· |
| T | 3808 hCoV-19/USA/CA-CZB-13558/2020\|EPI_ISL_672502\|2020- | ···················· | ·A·················· | ···················· | ···················· | ···················· | ···················· |
| T | 3810 hCoV-19/USA/CA-CZB-13530/2020\|EPI_ISL_672504\|2020- | ···················· | ···················· | ···················· | ···················· | ···················· | ···················· |
| T | 3811 hCoV-19/USA/CA-CZB-13528/2020\|EPI_ISL_672505\|2020- | ···················· | ···················· | ···················· | ···················· | ···················· | ···················· |
| T | 3812 hCoV-19/USA/CA-CZB-13527/2020\|EPI_ISL_672506\|2020- | ···················· | ·A·················· | ···················· | ···················· | ···················· | ···················· |
| T | 3814 hCoV-19/USA/CA-CZB-13522/2020\|EPI_ISL_672508\|2020- | ···················· | ·A·················· | ···················· | ···················· | ···················· | ···················· |
| T | 3815 hCoV-19/USA/CA-CZB-13520/2020\|EPI_ISL_672509\|2020- | ···················· | ·A·················· | ···················· | ···················· | ···················· | ···················· |
| T | 3816 hCoV-19/USA/CA-CZB-13519/2020\|EPI_ISL_672510\|2020- | ···················· | ·A·················· | ···················· | ···················· | ···················· | ···················· |
| T | 3817 hCoV-19/USA/CA-CZB-13514/2020\|EPI_ISL_672511\|2020- | ···················· | ···················· | ···················· | ···················· | ···················· | ···················· |
| T | 3818 hCoV-19/USA/CA-CZB-13513/2020\|EPI_ISL_672512\|2020- | ···················· | ···················· | ···················· | ···················· | ···················· | ···················· |
| T | 3819 hCoV-19/USA/CA-CZB-13511/2020\|EPI_ISL_672513\|2020- | ···················· | ·A·················· | ···················· | ···················· | ···················· | ···················· |
| T | 3821 hCoV-19/USA/CA-CZB-13507/2020\|EPI_ISL_672515\|2020- | ···················· | ·A·················· | ···················· | ···················· | ···················· | ···················· |
| T | 3822 hCoV-19/USA/CA-CZB-13506/2020\|EPI_ISL_672516\|2020- | ···················· | ·A·················· | ···················· | ···················· | ···················· | ···················· |
| T | 3823 hCoV-19/USA/CA-CZB-13505/2020\|EPI_ISL_672517\|2020- | ···················· | ·A·················· | ···················· | ···················· | ···················· | ···················· |
| T | 3824 hCoV-19/USA/CA-CZB-13504/2020\|EPI_ISL_672518\|2020- | ···················· | ·A·················· | ···················· | ···················· | ···················· | ···················· |
| T | 3825 hCoV-19/USA/CA-CZB-13502/2020\|EPI_ISL_672519\|2020- | ···················· | ···················· | ···················· | ···················· | ···················· | ···················· |
| T | 3826 hCoV-19/USA/CA-CZB-13500/2020\|EPI_ISL_672520\|2020- | ···················· | ·A·················· | ···················· | ···················· | ···················· | ···················· |
| T | 3827 hCoV-19/USA/CA-CZB-13498/2020\|EPI_ISL_672521\|2020- | ···················· | ·A·················· | ···················· | ···················· | ···················· | ···················· |
| T | 3828 hCoV-19/USA/CA-CZB-13489/2020\|EPI_ISL_672522\|2020- | ···················· | ···················· | ···················· | ···················· | ···················· | ···················· |
| T | 3829 hCoV-19/USA/CA-CZB-13487/2020\|EPI_ISL_672523\|2020- | ···················· | ·A·················· | ···················· | ···················· | ···················· | ···················· |
| T | 3830 hCoV-19/USA/CA-CZB-13486/2020\|EPI_ISL_672524\|2020- | ···················· | ···················· | ···················· | ···················· | ···················· | ···················· |
| T | 3831 hCoV-19/USA/CA-CZB-13479/2020\|EPI_ISL_672525\|2020- | ···················· | ·A·················· | ···················· | ···················· | ···················· | ···················· |
| T | 3832 hCoV-19/USA/CA-CZB-13477/2020\|EPI_ISL_672526\|2020- | ···················· | ···················· | ···················· | ···················· | ···················· | ···················· |
| T | 3833 hCoV-19/USA/CA-CZB-13476/2020\|EPI_ISL_672527\|2020- | ·······T············ | ···G················ | ···················· | ···················· | ···················· | ···················· |
| T | 3835 hCoV-19/USA/CA-CZB-13474/2020\|EPI_ISL_672529\|2020- | ·······T············ | ···G················ | ···················· | ···················· | ···················· | ···················· |
| T | 3837 hCoV-19/USA/UT-UPHL-201208035/2020\|EPI_ISL_672549\| | ···················· | ···················· | ···················· | ···················· | ···················· | ···················· |
| T | 3838 hCoV-19/USA/UT-UPHL-201225905/2020\|EPI_ISL_672550\| | ···················· | ···················· | ···················· | ···················· | ···················· | ···················· |
| T | 3839 hCoV-19/USA/UT-UPHL-201229235/2020\|EPI_ISL_672551\| | ···················· | ···················· | ···················· | ···················· | ···················· | ···················· |
| T | 3840 hCoV-19/USA/UT-UPHL-201227657/2020\|EPI_ISL_672552\| | ···················· | ···················· | ···················· | ···················· | ···················· | ···················· |
| T | 3841 hCoV-19/England/QEUH-B81075/2020\|EPI_ISL_661800\|20 | ···················· | ···················· | ···················· | ···················· | ···················· | ···················· |
| T | 3842 hCoV-19/England/CAMC-B785EB/2020\|EPI_ISL_661801\|20 | ···················· | ···················· | ···················· | ···················· | ······T············· | ···················· |
| T | 3843 hCoV-19/England/QEUH-B7F001/2020\|EPI_ISL_661802\|20 | ···················· | ···················· | ···················· | ···················· | ···················· | ···················· |
| T | 3844 hCoV-19/England/QEUH-B8286D/2020\|EPI_ISL_661803\|20 | ···················· | ·A·················· | ···················· | ···················· | ···················· | ··A················· |
| T | 3845 hCoV-19/England/QEUH-B80AE1/2020\|EPI_ISL_661804\|20 | ···················· | ···················· | ···················· | ···················· | ···················· | ···················· |
| T | 3846 hCoV-19/England/CAMC-B779E0/2020\|EPI_ISL_661805\|20 | ···················· | ···················· | ···················· | ···················· | ···················· | ···················· |
| T | 3847 hCoV-19/England/QEUH-B7F47E/2020\|EPI_ISL_661806\|20 | ···················· | ···················· | ···················· | ···················· | ···················· | ···················· |
| T | 3848 hCoV-19/England/QEUH-B80AD2/2020\|EPI_ISL_661807\|20 | ···················· | ···················· | ···················· | ···················· | ···················· | ···················· |
| T | 3849 hCoV-19/England/QEUH-B81312/2020\|EPI_ISL_661808\|20 | ···················· | ···················· | ···················· | ···················· | ···················· | ···················· |
| T | 3850 hCoV-19/England/QEUH-B7EF7E/2020\|EPI_ISL_661809\|20 | ···················· | ···················· | ···················· | ···················· | ···················· | ···················· |
| T | 3851 hCoV-19/England/QEUH-B7ED56/2020\|EPI_ISL_661810\|20 | ···················· | ···················· | ···················· | ···················· | ···················· | ···················· |
| T | 3852 hCoV-19/England/CAMC-B778A7/2020\|EPI_ISL_661811\|20 | ···················· | ···················· | ···················· | ···················· | ···················· | ···················· |
| T | 3853 hCoV-19/England/QEUH-B7EC3B/2020\|EPI_ISL_661812\|20 | ···················· | ···················· | ···················· | ···················· | ···················· | ···················· |
| T | 3854 hCoV-19/England/QEUH-B82645/2020\|EPI_ISL_661813\|20 | ···················· | ···················· | ···················· | ···················· | ···················· | ···················· |

T 3855 hCoV-19/England/QEUH-B7F3F9/2020|EPI_ISL_661816|20 ···················· ···················· ····················· ···················· ···················· ····················
T 3856 hCoV-19/England/CAMC-B77A0B/2020|EPI_ISL_661817|20 ···················· ···················· ····················· ···················· ·····T·············· ····················
T 3857 hCoV-19/England/QEUH-B81118/2020|EPI_ISL_661818|20 ···················· ···················· ····················· ···················· ·····T·············· ····················
T 3858 hCoV-19/England/QEUH-B7EB3E/2020|EPI_ISL_661819|20 ···················· ···················· ····················· ···················· ···················· ····················
T 3859 hCoV-19/England/QEUH-B804F2/2020|EPI_ISL_661823|20 ···················· ···················· ····················· ···················· ···················· ····················
T 3860 hCoV-19/England/QEUH-B805D1/2020|EPI_ISL_661824|20 ···················· ···················· ····················· ···················· ···················· ····················
T 3861 hCoV-19/England/QEUH-B80A69/2020|EPI_ISL_661825|20 ···················· ···················· ····················· ···················· ···················· ··A·················
T 3862 hCoV-19/England/QEUH-B7EF9C/2020|EPI_ISL_661827|20 ···················· ·A·················· ····················· ···················· ···················· ····················
T 3863 hCoV-19/England/QEUH-B807AE/2020|EPI_ISL_661828|20 ···················· ···················· ····················· ···················· ···················· ····················
T 3864 hCoV-19/England/QEUH-B80AA5/2020|EPI_ISL_661829|20 ···················· ···················· ····················· ···················· ···················· ····················
T 3865 hCoV-19/England/CAMC-B78581/2020|EPI_ISL_661830|20 ···················· ···················· ····················· ···················· ···················· ····················
T 3866 hCoV-19/England/QEUH-B7E406/2020|EPI_ISL_661831|20 ···················· ···················· ····················· ···················· ···················· ····················
T 3867 hCoV-19/England/QEUH-B809C6/2020|EPI_ISL_661832|20 ···················· ···················· ····················· ···················· ···················· ····················
T 3868 hCoV-19/England/CAMC-B77546/2020|EPI_ISL_661833|20 ···················· ···················· ····················· ···················· ···················· ····················
T 3869 hCoV-19/England/QEUH-B80C63/2020|EPI_ISL_661835|20 ···················· ···················· ····················· ···················· ···················· ····················
T 3870 hCoV-19/England/CAMC-B782E4/2020|EPI_ISL_661836|20 ···················· ···················· ····················· ···················· ···················· ····················
T 3871 hCoV-19/England/QEUH-B7EF50/2020|EPI_ISL_661837|20 ···················· ···················· ····················· ···················· ···················· ····················
T 3872 hCoV-19/England/QEUH-B82241/2020|EPI_ISL_661839|20 ···················· ···················· ····················· ···················· ·····T·············· ····················
T 3873 hCoV-19/England/ALDP-B74DE7/2020|EPI_ISL_662214|20 ···················· ···················· ····················· ···················· ···················· ····················
T 3874 hCoV-19/England/ALDP-B75557/2020|EPI_ISL_662215|20 ···················· ···················· ····················· ···················· ···················· ····················
T 3875 hCoV-19/England/QEUH-B66E7C/2020|EPI_ISL_662216|20 ···················· ···················· ····················· ···················· ···················· ····················
T 3876 hCoV-19/England/ALDP-B74646/2020|EPI_ISL_662217|20 ···················· ···················· ····················· ···················· ···················· ····················
T 3877 hCoV-19/England/ALDP-B7445B/2020|EPI_ISL_662218|20 ···················· ···················· ····················· ···················· ···················· ····················
T 3878 hCoV-19/England/ALDP-B75180/2020|EPI_ISL_662219|20 ···················· ···················· ····················· ···················· ···················· ····················
T 3879 hCoV-19/England/QEUH-B6746A/2020|EPI_ISL_662220|20 ···················· ···················· ····················· ···················· ···················· ····················
T 3880 hCoV-19/England/QEUH-B67F96/2020|EPI_ISL_662221|20 ···················· ···················· ····················· ···················· ···················· ····················
T 3881 hCoV-19/England/QEUH-B69107/2020|EPI_ISL_662225|20 ···················· ···················· ····················· ···················· ···················· ····················
T 3882 hCoV-19/England/QEUH-B69240/2020|EPI_ISL_662226|20 ···················· ·A·················· ····················· ···················· ···················· ··A·················
T 3883 hCoV-19/England/ALDP-B74691/2020|EPI_ISL_662227|20 ···················· ···················· ····················· ···················· ···················· ····················
T 3884 hCoV-19/England/ALDP-B74C53/2020|EPI_ISL_662228|20 ···················· ···················· ····················· ···················· ···················· ····················
T 3885 hCoV-19/England/ALDP-B75575/2020|EPI_ISL_662229|20 ···················· ···················· ····················· ···················· ···················· ····················
T 3886 hCoV-19/England/QEUH-B691F8/2020|EPI_ISL_662230|20 ···················· ···················· ····················· ···················· ···················· ····················
T 3887 hCoV-19/England/ALDP-B74057/2020|EPI_ISL_662231|20 ···················· ···················· ····················· ···················· ···················· ····················
T 3888 hCoV-19/England/QEUH-B68A1C/2020|EPI_ISL_662232|20 ···················· ···················· ····················· ···················· ···················· ····················
T 3889 hCoV-19/England/ALDP-B74734/2020|EPI_ISL_662233|20 ···················· ···················· ····················· ···················· ···················· ····················
T 3890 hCoV-19/England/ALDP-B75311/2020|EPI_ISL_662235|20 ···················· ···················· ····················· ···················· ···················· ····················
T 3891 hCoV-19/England/ALDP-B752D8/2020|EPI_ISL_662237|20 ···················· ···················· ····················· ···················· ···················· ····················
T 3892 hCoV-19/England/ALDP-B75074/2020|EPI_ISL_662238|20 ···················· ···················· ····················· ···················· ···················· ····················
T 3893 hCoV-19/England/ALDP-B73FC4/2020|EPI_ISL_662239|20 ···················· ···················· ····················· ···················· ···················· ····················
T 3894 hCoV-19/England/ALDP-B749C5/2020|EPI_ISL_662240|20 ···················· ···················· ····················· ···················· ···················· ····················
T 3895 hCoV-19/England/QEUH-B68DC8/2020|EPI_ISL_662241|20 ···················· ···················· ····················· ···················· ···················· ····················
T 3896 hCoV-19/England/QEUH-B68056/2020|EPI_ISL_662243|20 ···················· ···················· ····················· ···················· ···················· ····················
T 3897 hCoV-19/England/ALDP-B742E8/2020|EPI_ISL_662244|20 ···················· ···················· ····················· ···················· ···················· ····················
T 3898 hCoV-19/England/ALDP-B7526F/2020|EPI_ISL_662245|20 ···················· ···················· ····················· ···················· ···················· ····················
T 3899 hCoV-19/England/QEUH-B67FE1/2020|EPI_ISL_662246|20 ···················· ···················· ····················· ···················· ···················· ····················
T 3900 hCoV-19/England/ALDP-B75812/2020|EPI_ISL_662247|20 ···················· ···················· ····················· ···················· ···················· ····················
T 3901 hCoV-19/England/ALDP-B74C62/2020|EPI_ISL_662248|20 ···················· ···················· ····················· ···················· ···················· ····················
T 3902 hCoV-19/England/ALDP-B757CA/2020|EPI_ISL_662249|20 ···················· ···················· ····················· ···················· ···················· ····················
T 3903 hCoV-19/England/QEUH-B673B8/2020|EPI_ISL_662250|20 ···················· ···················· ····················· ···················· ···················· ····················
T 3904 hCoV-19/England/ALDP-B750A1/2020|EPI_ISL_662252|20 ···················· ···················· ····················· ···················· ···················· ····················
T 3905 hCoV-19/England/QEUH-B6743D/2020|EPI_ISL_662253|20 ···················· ···················· ····················· ···················· ···················· ····················
T 3906 hCoV-19/Wales/ALDP-B74D50/2020|EPI_ISL_662254|2020 ···················· ···················· ····················· ···················· ···················· ····················
T 3907 hCoV-19/England/ALDP-B7401B/2020|EPI_ISL_662255|20 ···················· ···················· ····················· ···················· ···················· ····················
T 3908 hCoV-19/England/QEUH-B67FF0/2020|EPI_ISL_662256|20 ···················· ···················· ····················· ···················· ···················· ····················
T 3910 hCoV-19/Australia/VIC15561/2020|EPI_ISL_663547|202 ···················· ···················· ····················· ···················· ···················· ····················
T 3911 hCoV-19/Australia/VIC15572/2020|EPI_ISL_663548|202 ···················· ···················· ····················· ···················· ···················· ····················
T 3917 hCoV-19/Australia/VIC16731/2020|EPI_ISL_663556|202 ···················· ···················· ····················· ···················· ···················· ····················
T 3918 hCoV-19/Australia/VIC16732/2020|EPI_ISL_663557|202 ···················· ···················· ····················· ···················· ···················· ····················
T 3919 hCoV-19/Australia/VIC16733/2020|EPI_ISL_663558|202 ···················· ···················· ····················· ···················· ···················· ····················
T 3920 hCoV-19/Australia/VIC16734/2020|EPI_ISL_663559|202 ···················· ···················· ····················· ···················· ···················· ····················
T 3923 hCoV-19/Australia/VIC16742/2020|EPI_ISL_663562|202 ···················· ···················· ····················· ···················· ···················· ····················
T 3929 hCoV-19/Australia/VIC16753/2020|EPI_ISL_663568|202 ···················· ···················· ····················· ···················· ···················· ····················
T 3933 hCoV-19/Australia/VIC16765/2020|EPI_ISL_663572|202 ···················· ···················· ····················· ···················· ···················· ····················
T 3934 hCoV-19/Australia/VIC16767/2020|EPI_ISL_663573|202 ···················· ···················· ····················· ···················· ···················· ····················
T 3935 hCoV-19/Australia/VIC16771/2020|EPI_ISL_663574|202 ···················· ···················· ····················· ···················· ···················· ····················
T 3936 hCoV-19/Australia/VIC16772/2020|EPI_ISL_663575|202 ···················· ···················· ····················· ···················· ···················· ····················
T 3937 hCoV-19/Australia/VIC16774/2020|EPI_ISL_663576|202 ···················· ···················· ····················· ···················· ···················· ····················
T 3938 hCoV-19/Australia/VIC16780/2020|EPI_ISL_663577|202 ···················· ···················· ····················· ···················· ···················· ····················
T 3939 hCoV-19/Australia/VIC16782/2020|EPI_ISL_663578|202 ···················· ···················· ····················· ···················· ···················· ····················
T 3940 hCoV-19/Australia/VIC16785/2020|EPI_ISL_663579|202 ···················· ···················· ····················· ···················· ···················· ····················
T 3941 hCoV-19/Australia/VIC16786/2020|EPI_ISL_663580|202 ···················· ···················· ····················· ···················· ···················· ····················
T 3942 hCoV-19/Australia/VIC16787/2020|EPI_ISL_663581|202 ···················· ···················· ····················· ···················· ···················· ····················
T 3943 hCoV-19/Australia/VIC16796/2020|EPI_ISL_663582|202 ···················· ···················· ····················· ···················· ···················· ····················
T 3945 hCoV-19/Australia/VIC16799/2020|EPI_ISL_663584|202 ···················· ···················· ····················· ···················· ···················· ····················
T 3946 hCoV-19/Australia/VIC16800/2020|EPI_ISL_663585|202 ···················· ···················· ····················· ···················· ···················· ····················
T 3947 hCoV-19/Australia/VIC16818/2020|EPI_ISL_663586|202 ···················· ······A············· ····················· ···················· ···················· ····················
T 3948 hCoV-19/Australia/VIC16821/2020|EPI_ISL_663587|202 ···················· ···················· ····················· ···················· ···················· ····················
T 3949 hCoV-19/England/C200044586/2020|EPI_ISL_664043|202 ···················· ···················· ····················· ···················· ···················· ····················
T 3950 hCoV-19/England/C200045611/2020|EPI_ISL_664055|202 ···················· ·········T·········· ····················· ···················· ···················· ····················
T 3951 hCoV-19/England/C200045757/2020|EPI_ISL_664070|202 ···················· ···················· ····················· ···················· ···················· ····················

| | | | | | | | | |
|---|---|---|---|---|---|---|---|---|
| T | 3952 | hCoV-19/England/C200045797/2020\|EPI_ISL_664080\|202 | ···················· | ···················· | ····················· | ···················· | ···················· | ···················· |
| T | 3953 | hCoV-19/England/C200045839/2020\|EPI_ISL_664084\|202 | ···················· | ···················· | ····················· | ···················· | ···················· | ···················· |
| T | 3954 | hCoV-19/England/ALDP-B73C36/2020\|EPI_ISL_664875\|20 | ···················· | ···················· | ····················· | ···················· | ···················· | ···················· |
| T | 3955 | hCoV-19/England/ALDP-B73C45/2020\|EPI_ISL_664876\|20 | ···················· | ···················· | ····················· | ···················· | ···················· | ···················· |
| T | 3956 | hCoV-19/England/ALDP-B73C54/2020\|EPI_ISL_664877\|20 | ···················· | ···················· | ····················· | ···················· | ···················· | ···················· |
| T | 3957 | hCoV-19/England/ALDP-B73C72/2020\|EPI_ISL_664878\|20 | ···················· | ···················· | ····················· | ···················· | ···················· | ···················· |
| T | 3958 | hCoV-19/England/ALDP-B73C81/2020\|EPI_ISL_664879\|20 | ···················· | ···················· | ····················· | ···················· | ···················· | ···················· |
| T | 3959 | hCoV-19/England/ALDP-B73C90/2020\|EPI_ISL_664880\|20 | ···················· | ···················· | ····················· | ···················· | ···················· | ···················· |
| T | 3960 | hCoV-19/England/ALDP-B73CCD/2020\|EPI_ISL_664881\|20 | ···················· | ···················· | ····················· | ···················· | ···················· | ···················· |
| T | 3962 | hCoV-19/England/ALDP-B73D15/2020\|EPI_ISL_664883\|20 | ···················· | ···················· | ····················· | ···················· | ···················· | ···················· |
| T | 3963 | hCoV-19/England/ALDP-B73D33/2020\|EPI_ISL_664885\|20 | ···················· | ···················· | ····················· | ···················· | ···················· | ···················· |
| T | 3965 | hCoV-19/England/ALDP-B73D9D/2020\|EPI_ISL_664887\|20 | ···················· | ···················· | ····················· | ···················· | ···················· | ···················· |
| T | 3969 | hCoV-19/England/ALDP-B73DD9/2020\|EPI_ISL_664891\|20 | ···················· | ···················· | ····················· | ···················· | ···················· | ···················· |
| T | 3970 | hCoV-19/England/ALDP-B73E03/2020\|EPI_ISL_664892\|20 | ···················· | ·A·················· | ····················· | ···················· | ···················· | ···················· |
| T | 3971 | hCoV-19/England/ALDP-B73E21/2020\|EPI_ISL_664893\|20 | ···················· | ···················· | ····················· | ···················· | ···················· | ···················· |
| T | 3972 | hCoV-19/England/ALDP-B73E5E/2020\|EPI_ISL_664894\|20 | ···················· | ·A·················· | ····················· | ···················· | ···················· | ···A················ |
| T | 3973 | hCoV-19/England/ALDP-B73E8B/2020\|EPI_ISL_664895\|20 | ···················· | ···················· | ····················· | ···················· | ···················· | ···················· |
| T | 3974 | hCoV-19/England/ALDP-B73EB8/2020\|EPI_ISL_664896\|20 | ···················· | ···················· | ····················· | ···················· | ···················· | ···················· |
| T | 3975 | hCoV-19/England/ALDP-B73ED6/2020\|EPI_ISL_664897\|20 | ···················· | ···················· | ····················· | ···················· | ···················· | ···················· |
| T | 3976 | hCoV-19/England/ALDP-B73F88/2020\|EPI_ISL_664898\|20 | ···················· | ···················· | ····················· | ···················· | ···················· | ···················· |
| T | 3977 | hCoV-19/England/ALDP-B74039/2020\|EPI_ISL_664899\|20 | ···················· | ···················· | ····················· | ···················· | ···················· | ···················· |
| T | 3978 | hCoV-19/USA/LA-EVTL1031/2020\|EPI_ISL_666613\|2020-0 | ···················· | ···················· | ····················· | ···················· | ···················· | ···················· |
| T | 3979 | hCoV-19/USA/FL-UF-31/2020\|EPI_ISL_666617\|2020-09-2 | ···················· | ···················· | ····················· | ···················· | ···················· | ···················· |
| T | 3980 | hCoV-19/USA/FL-UF-32/2020\|EPI_ISL_666618\|2020-10-1 | ···················· | ·A·················· | ····················· | ···················· | ···················· | ···················· |
| T | 3981 | hCoV-19/USA/FL-UF-33/2020\|EPI_ISL_666619\|2020-10-1 | ···················· | ·A·················· | ····················· | ···················· | ···················· | ···················· |
| T | 3982 | hCoV-19/USA/FL-UF-34/2020\|EPI_ISL_666620\|2020-10-1 | ···················· | ·A·················· | ····················· | ···················· | ···················· | ···················· |
| T | 3983 | hCoV-19/USA/FL-UF-35/2020\|EPI_ISL_666621\|2020-10-1 | ···················· | ·A·················· | ····················· | ···················· | ···················· | ···················· |
| T | 3984 | hCoV-19/USA/FL-UF-36/2020\|EPI_ISL_666622\|2020-10-0 | ···················· | ·A·················· | ····················· | ···················· | ···················· | ···················· |
| T | 3985 | hCoV-19/Germany/NW-HHU-321/2020\|EPI_ISL_666623\|202 | ···················· | ···················· | ····················· | ···················· | ···················· | ···················· |
| T | 3986 | hCoV-19/Germany/NW-HHU-324/2020\|EPI_ISL_666625\|202 | ···················· | ···················· | ····················· | ···················· | ···················· | ···················· |
| T | 3987 | hCoV-19/Germany/NW-HHU-334/2020\|EPI_ISL_666628\|202 | ···················· | ···················· | ····················· | ···················· | ···················· | ···················· |
| T | 3988 | hCoV-19/Germany/NW-HHU-340/2020\|EPI_ISL_666629\|202 | ···················· | ···················· | ····················· | ···················· | ···················· | ···················· |
| T | 3989 | hCoV-19/USA/CA-ALSR-4598/2020\|EPI_ISL_667030\|2020- | ···················· | ···················· | ····················· | ···················· | ···················· | ···················· |
| T | 3990 | hCoV-19/USA/CA-ALSR-4599/2020\|EPI_ISL_667031\|2020- | ···················· | ·A·················· | ····················· | ···················· | ···················· | ···················· |
| T | 3991 | hCoV-19/USA/CA-ALSR-4602/2020\|EPI_ISL_667032\|2020- | ···················· | ···················· | ····················· | ···················· | ···················· | ···················· |
| T | 3992 | hCoV-19/USA/CA-ALSR-4604/2020\|EPI_ISL_667033\|2020- | ···················· | ·A·················· | ····················· | ···················· | ···················· | ···················· |
| T | 3993 | hCoV-19/USA/CA-ALSR-4607/2020\|EPI_ISL_667036\|2020- | ···················· | ·A·················· | ····················· | ···················· | ···················· | ···················· |
| T | 3994 | hCoV-19/USA/CA-ALSR-4608/2020\|EPI_ISL_667037\|2020- | ···················· | ·A·················· | ····················· | ···················· | ···················· | ···················· |
| T | 3995 | hCoV-19/USA/CA-ALSR-4611/2020\|EPI_ISL_667038\|2020- | ···················· | ·A·················· | ····················· | ···················· | ···················· | ···················· |
| T | 3996 | hCoV-19/USA/CA-ALSR-4613/2020\|EPI_ISL_667039\|2020- | ···················· | ·A·················· | ····················· | ···················· | ···················· | ···················· |
| T | 3997 | hCoV-19/USA/CA-ALSR-4616/2020\|EPI_ISL_667040\|2020- | ···················· | ·A·················· | ····················· | ···················· | ···················· | ···················· |
| T | 3998 | hCoV-19/USA/CA-ALSR-4619/2020\|EPI_ISL_667041\|2020- | ···················· | ·A·················· | ····················· | ···················· | ···················· | ···················· |
| T | 3999 | hCoV-19/USA/CA-ALSR-4622/2020\|EPI_ISL_667042\|2020- | ···················· | ···················· | ····················· | ···················· | ···················· | ···················· |
| T | 4000 | hCoV-19/USA/CA-ALSR-4623/2020\|EPI_ISL_667043\|2020- | ···················· | ·A·················· | ····················· | ···················· | ···················· | ···················· |
| T | 4001 | hCoV-19/USA/CA-ALSR-4624/2020\|EPI_ISL_667044\|2020- | ···················· | ·············A······ | ····················· | ···················· | ···················· | ···················· |
| T | 4002 | hCoV-19/USA/CA-ALSR-4625/2020\|EPI_ISL_667045\|2020- | ···················· | ·A·················· | ····················· | ···················· | ···················· | ···················· |
| T | 4003 | hCoV-19/USA/CA-ALSR-4628/2020\|EPI_ISL_667046\|2020- | ···················· | ·A·················· | ····················· | ···················· | ···················· | ···················· |
| T | 4004 | hCoV-19/USA/CA-ALSR-4629/2020\|EPI_ISL_667047\|2020- | ···················· | ·A·················· | ····················· | ···················· | ···················· | ···················· |
| T | 4005 | hCoV-19/USA/CA-ALSR-4633/2020\|EPI_ISL_667048\|2020- | ···················· | ·A·················· | ····················· | ···················· | ···················· | ···················· |
| T | 4006 | hCoV-19/USA/CA-ALSR-4636/2020\|EPI_ISL_667049\|2020- | ···················· | ·A·················· | ····················· | ···················· | ···················· | ···················· |
| T | 4007 | hCoV-19/USA/CA-ALSR-4637/2020\|EPI_ISL_667050\|2020- | ···················· | ·A·················· | ····················· | ···················· | ···················· | ···················· |
| T | 4008 | hCoV-19/USA/CA-ALSR-4639/2020\|EPI_ISL_667051\|2020- | ···················· | ···················· | ····················· | ···················· | ···················· | ···············A···· |
| T | 4009 | hCoV-19/USA/CA-ALSR-4640/2020\|EPI_ISL_667052\|2020- | ···················· | ·A·················· | ····················· | ···················· | ···················· | ···················· |
| T | 4010 | hCoV-19/USA/CA-ALSR-4644/2020\|EPI_ISL_667054\|2020- | ···················· | ·A·················· | ····················· | ···················· | ···················· | ···················· |
| T | 4011 | hCoV-19/USA/CA-ALSR-4646/2020\|EPI_ISL_667055\|2020- | ···················· | ·A·················· | ····················· | ···················· | ···················· | ···················· |
| T | 4012 | hCoV-19/USA/CA-ALSR-4649/2020\|EPI_ISL_667056\|2020- | ···················· | ·A·················· | ····················· | ···················· | ···················· | ···················· |
| T | 4013 | hCoV-19/USA/CA-ALSR-4651/2020\|EPI_ISL_667057\|2020- | ···················· | ···················· | ····················· | ···················· | ···················· | ···················· |
| T | 4014 | hCoV-19/USA/CA-ALSR-4652/2020\|EPI_ISL_667058\|2020- | ···················· | ·A·················· | ····················· | ···················· | ···················· | ···················· |
| T | 4015 | hCoV-19/USA/CA-ALSR-4654/2020\|EPI_ISL_667059\|2020- | ···················· | ·A·················· | ····················· | ···················· | ···················· | ···················· |
| T | 4016 | hCoV-19/USA/CA-ALSR-4658/2020\|EPI_ISL_667060\|2020- | ···················· | ·A·················· | ····················· | ···················· | ···················· | ···················· |
| T | 4017 | hCoV-19/USA/CA-ALSR-4660/2020\|EPI_ISL_667061\|2020- | ···················· | ·A·················· | ····················· | ···················· | ···················· | ···················· |
| T | 4018 | hCoV-19/USA/CA-ALSR-4662/2020\|EPI_ISL_667062\|2020- | ···················· | ·A·················· | ····················· | ···················· | ···················· | ···················· |
| T | 4019 | hCoV-19/USA/CA-ALSR-4663/2020\|EPI_ISL_667063\|2020- | ···················· | ·A·················· | ····················· | ···················· | ···················· | ···················· |
| T | 4020 | hCoV-19/USA/CA-ALSR-4664/2020\|EPI_ISL_667064\|2020- | ···················· | ···················· | ····················· | ···················· | ···················· | ···················· |
| T | 4021 | hCoV-19/USA/CA-ALSR-4665/2020\|EPI_ISL_667065\|2020- | ···················· | ·A·················· | ····················· | ···················· | ···················· | ···················· |
| T | 4022 | hCoV-19/USA/CA-ALSR-4666/2020\|EPI_ISL_667066\|2020- | ···················· | ·A·················· | ····················· | ···················· | ···················· | ···················· |
| T | 4023 | hCoV-19/USA/CA-ALSR-4669/2020\|EPI_ISL_667067\|2020- | ···················· | ···················· | ····················· | ···················· | ···················· | ···················· |
| T | 4024 | hCoV-19/USA/CA-ALSR-4670/2020\|EPI_ISL_667068\|2020- | ···················· | ·A·················· | ····················· | ···················· | ···················· | ···················· |
| T | 4025 | hCoV-19/USA/CA-ALSR-4672/2020\|EPI_ISL_667069\|2020- | ···················· | ·A·················· | ····················· | ···················· | ···················· | ···················· |
| T | 4026 | hCoV-19/USA/CA-ALSR-4674/2020\|EPI_ISL_667070\|2020- | ···················· | ·A·················· | ····················· | ···················· | ···················· | ···················· |
| T | 4027 | hCoV-19/USA/CA-ALSR-4676/2020\|EPI_ISL_667072\|2020- | ···················· | ·A·················· | ····················· | ···················· | ···················· | ···················· |
| T | 4028 | hCoV-19/USA/CA-ALSR-4677/2020\|EPI_ISL_667073\|2020- | ···················· | ···················· | ····················· | ···················· | ···················· | ···················· |
| T | 4029 | hCoV-19/USA/CA-ALSR-4682/2020\|EPI_ISL_667074\|2020- | ···················· | ·A·················· | ····················· | ···················· | ···················· | ···················· |
| T | 4030 | hCoV-19/USA/CA-ALSR-4683/2020\|EPI_ISL_667075\|2020- | ···················· | ···················· | ····················· | ···················· | ···················· | ···················· |
| T | 4031 | hCoV-19/USA/CA-ALSR-4685/2020\|EPI_ISL_667076\|2020- | ···················· | ·A·················· | ····················· | ···················· | ···················· | ···················· |
| T | 4032 | hCoV-19/USA/CA-ALSR-4686/2020\|EPI_ISL_667077\|2020- | ···················· | ···················· | ····················· | ···················· | ···················· | ···················· |
| T | 4033 | hCoV-19/USA/OR-OHSU-0070/2020\|EPI_ISL_667079\|2020- | ···················· | ···················· | ····················· | ···················· | ···················· | ···················· |
| T | 4034 | hCoV-19/USA/OR-OHSU-1886/2020\|EPI_ISL_667085\|2020- | ···················· | ···················· | ····················· | ···················· | ···················· | ···················· |
| T | 4035 | hCoV-19/USA/OR-OHSU-1972/2020\|EPI_ISL_667086\|2020- | ···················· | ···················· | ····················· | ···················· | ···················· | ···················· |
| T | 4036 | hCoV-19/USA/OR-OHSU-2003/2020\|EPI_ISL_667087\|2020- | ···················· | ···················· | ····················· | ···················· | ···················· | ···················· |

| | | | | | | | |
|---|---|---|---|---|---|---|---|
| T | 4037 hCoV-19/USA/CA-OHSU-2007/2020\|EPI_ISL_667088\|2020- | ···················· | ···················· | ····················· | ···················· | ···················· | ···················· |
| T | 4038 hCoV-19/USA/OR-OHSU-2009/2020\|EPI_ISL_667089\|2020- | ···················· | ···················· | ····················· | ···················· | ···················· | ···················· |
| T | 4039 hCoV-19/USA/OR-OHSU-2011/2020\|EPI_ISL_667090\|2020- | ···················· | ···················· | ····················· | ···················· | ···················· | ···················· |
| T | 4040 hCoV-19/USA/OR-OHSU-2015/2020\|EPI_ISL_667091\|2020- | ···················· | ···················· | ····················· | ···················· | ···················· | ···················· |
| T | 4041 hCoV-19/USA/OR-OHSU-2017/2020\|EPI_ISL_667092\|2020- | ···················· | ···················· | ····················· | ···················· | ···················· | ···················· |
| T | 4042 hCoV-19/USA/OR-OHSU-2023/2020\|EPI_ISL_667093\|2020- | ···················· | ···················· | ····················· | ···················· | ···················· | ···················· |
| T | 4043 hCoV-19/USA/OR-OHSU-2025/2020\|EPI_ISL_667094\|2020- | ···················· | ···················· | ····················· | ···················· | ···················· | ···················· |
| T | 4044 hCoV-19/USA/OR-OHSU-2027/2020\|EPI_ISL_667095\|2020- | ···················· | ···················· | ····················· | ···················· | ···················· | ···················· |
| T | 4045 hCoV-19/USA/OR-OHSU-2029/2020\|EPI_ISL_667096\|2020- | ···················· | ···················· | ····················· | ···················· | ···················· | ···················· |
| T | 4046 hCoV-19/USA/WA-OHSU-2033/2020\|EPI_ISL_667098\|2020- | ···················· | ···················· | ····················· | ···················· | ···················· | ···················· |
| T | 4047 hCoV-19/USA/OR-OHSU-2035/2020\|EPI_ISL_667099\|2020- | ···················· | ···················· | ····················· | ···················· | ···················· | ···················· |
| T | 4048 hCoV-19/England/CAMC-B7968D/2020\|EPI_ISL_667944\|20 | ···················· | ···················· | ····················· | ···················· | ···················· | ···················· |
| T | 4049 hCoV-19/England/CAMC-B7AE57/2020\|EPI_ISL_667945\|20 | ···················· | ···················· | ····················· | ···················· | ·····T·············· | ···············A···· |
| T | 4050 hCoV-19/England/CAMC-B7B0E7/2020\|EPI_ISL_667946\|20 | ···················· | ·A·················· | ····················· | ···················· | ···················· | ···A················ |
| T | 4051 hCoV-19/England/QEUH-B80FA6/2020\|EPI_ISL_667947\|20 | ···················· | ···················· | ····················· | ···················· | ···················· | ···················· |
| T | 4052 hCoV-19/England/QEUH-B7E54F/2020\|EPI_ISL_667950\|20 | ···················· | ···················· | ····················· | ···················· | ···················· | ···················· |
| T | 4053 hCoV-19/England/QEUH-B7E521/2020\|EPI_ISL_667952\|20 | ···················· | ···················· | ····················· | ···················· | ···················· | ···················· |
| T | 4054 hCoV-19/England/CAMC-B799B1/2020\|EPI_ISL_667953\|20 | ···················· | ···················· | ····················· | ···················· | ···················· | ···················· |
| T | 4055 hCoV-19/England/CAMC-B7AA08/2020\|EPI_ISL_667954\|20 | ···················· | ···················· | ····················· | ···················· | ···················· | ···················· |
| T | 4056 hCoV-19/England/QEUH-B7EE35/2020\|EPI_ISL_667959\|20 | ···················· | ···················· | ····················· | ···················· | ···················· | ···················· |
| T | 4057 hCoV-19/England/QEUH-B7F1FF/2020\|EPI_ISL_667960\|20 | ···················· | ···················· | ····················· | ···················· | ···················· | ···················· |
| T | 4058 hCoV-19/England/QEUH-B81B47/2020\|EPI_ISL_667962\|20 | ···················· | ···················· | ····················· | ···················· | ···················· | ···················· |
| T | 4059 hCoV-19/England/QEUH-B7E57C/2020\|EPI_ISL_667963\|20 | ···················· | ···················· | ····················· | ···················· | ···················· | ···················· |
| T | 4060 hCoV-19/England/CAMC-B7B09C/2020\|EPI_ISL_667965\|20 | ···················· | ···················· | ····················· | ···················· | ···················· | ···················· |
| T | 4061 hCoV-19/England/QEUH-B806ED/2020\|EPI_ISL_668360\|20 | ···················· | ·A·················· | ····················· | ···················· | ···················· | ···················· |
| T | 4062 hCoV-19/England/QEUH-B7F98E/2020\|EPI_ISL_668362\|20 | ···················· | ···················· | ····················· | ···················· | ···················· | ···················· |
| T | 4063 hCoV-19/England/QEUH-B7E3CD/2020\|EPI_ISL_668364\|20 | ···················· | ···················· | ····················· | ···················· | ···················· | ···················· |
| T | 4064 hCoV-19/England/QEUH-B8250C/2020\|EPI_ISL_668366\|20 | ···················· | ···················· | ····················· | ···················· | ·····T·············· | ···················· |
| T | 4065 hCoV-19/England/QEUH-B80638/2020\|EPI_ISL_668367\|20 | ···················· | ···················· | ····················· | ···················· | ···················· | ···················· |
| T | 4066 hCoV-19/England/QEUH-B7F9D9/2020\|EPI_ISL_668372\|20 | ···················· | ···················· | ····················· | ···················· | ·····T·············· | ···················· |
| T | 4067 hCoV-19/England/CAMC-B78123/2020\|EPI_ISL_668373\|20 | ···················· | ···················· | ····················· | ···················· | ···················· | ···················· |
| T | 4068 hCoV-19/England/CAMC-B77FC0/2020\|EPI_ISL_668374\|20 | ···················· | ···················· | ····················· | ···················· | ···················· | ···················· |
| T | 4069 hCoV-19/England/QEUH-B80656/2020\|EPI_ISL_668380\|20 | ···················· | ·A·················· | ····················· | ···················· | ···················· | ···················· |
| T | 4070 hCoV-19/England/QEUH-B7F5D5/2020\|EPI_ISL_668381\|20 | ···················· | ···················· | ····················· | ···················· | ···················· | ···················· |
| T | 4071 hCoV-19/England/QEUH-B7C3FC/2020\|EPI_ISL_668383\|20 | ···················· | ···················· | ····················· | ···················· | ···················· | ···················· |
| T | 4072 hCoV-19/England/QEUH-B7C12F/2020\|EPI_ISL_668386\|20 | ···················· | ···················· | ····················· | ···················· | ···················· | ···················· |
| T | 4073 hCoV-19/England/QEUH-B7FD73/2020\|EPI_ISL_668387\|20 | ···················· | ···················· | ····················· | ···················· | ···T················ | ···················· |
| T | 4074 hCoV-19/England/CAMC-B78044/2020\|EPI_ISL_668388\|20 | ···················· | ···················· | ····················· | ···················· | ···················· | ···················· |
| T | 4075 hCoV-19/England/CAMC-B7785C/2020\|EPI_ISL_668389\|20 | ···················· | ·A·················· | ····················· | ···················· | ···················· | ···················· |
| T | 4076 hCoV-19/Norway/4686/2020\|EPI_ISL_668393\|2020-09-30 | ···················· | ···················· | ····················· | ···················· | ···················· | ···················· |
| T | 4077 hCoV-19/Norway/4724/2020\|EPI_ISL_668394\|2020-10-21 | ···················· | ···················· | ····················· | ···················· | ···················· | ···················· |
| T | 4078 hCoV-19/Norway/4725/2020\|EPI_ISL_668395\|2020-10-21 | ···················· | ·A·················· | ····················· | ···················· | ···················· | ···A················ |
| T | 4079 hCoV-19/Norway/4749/2020\|EPI_ISL_668396\|2020-10-22 | ···················· | ···················· | ····················· | ···················· | ···················· | ···················· |
| T | 4080 hCoV-19/Norway/4752/2020\|EPI_ISL_668397\|2020-10-22 | ···················· | ···················· | ····················· | ···················· | ···················· | ···················· |
| T | 4081 hCoV-19/Norway/4755/2020\|EPI_ISL_668398\|2020-10-13 | ···················· | ···················· | ····················· | ···················· | ····C··············· | ···················· |
| T | 4082 hCoV-19/Denmark/DCGC-10784/2020\|EPI_ISL_669690\|202 | ···················· | ···················· | ····················· | ···················· | ···················· | ···················· |
| T | 4083 hCoV-19/Denmark/DCGC-10802/2020\|EPI_ISL_669691\|202 | ···················· | ···················· | ····················· | ···················· | ···················· | ···················· |
| T | 4084 hCoV-19/Denmark/DCGC-10808/2020\|EPI_ISL_669692\|202 | ···················· | ···················· | ····················· | ···················· | ···················· | ···················· |
| T | 4085 hCoV-19/Denmark/DCGC-10810/2020\|EPI_ISL_669693\|202 | ···················· | ···················· | ····················· | ···················· | ···················· | ···················· |
| T | 4086 hCoV-19/Denmark/DCGC-10842/2020\|EPI_ISL_669694\|202 | ···················· | ···················· | ····················· | ···················· | ···················· | ···················· |
| T | 4087 hCoV-19/Denmark/DCGC-10843/2020\|EPI_ISL_669695\|202 | ···················· | ···················· | ····················· | ···················· | ···················· | ···················· |
| T | 4088 hCoV-19/Denmark/DCGC-10850/2020\|EPI_ISL_669696\|202 | ···················· | ···················· | ····················· | ···················· | ···················· | ···················· |
| T | 4089 hCoV-19/Denmark/DCGC-10856/2020\|EPI_ISL_669697\|202 | ···················· | ···················· | ····················· | ···················· | ···················· | ···················· |
| T | 4090 hCoV-19/Denmark/DCGC-10863/2020\|EPI_ISL_669698\|202 | ···················· | ···················· | ····················· | ···················· | ···················· | ···················· |
| T | 4091 hCoV-19/Denmark/DCGC-10874/2020\|EPI_ISL_669699\|202 | ···················· | ···················· | ····················· | ···················· | ···················· | ···················· |
| T | 4092 hCoV-19/Denmark/DCGC-11966/2020\|EPI_ISL_670672\|202 | ···················· | ···················· | ····················· | ···················· | ···················· | ···················· |
| T | 4093 hCoV-19/Denmark/DCGC-11967/2020\|EPI_ISL_670673\|202 | ···················· | ···················· | ····················· | ···················· | ···················· | ···················· |
| T | 4094 hCoV-19/Denmark/DCGC-11973/2020\|EPI_ISL_670674\|202 | ···················· | ···················· | ····················· | ···················· | ···················· | ···················· |
| T | 4095 hCoV-19/Denmark/DCGC-11554/2020\|EPI_ISL_670680\|202 | ···················· | ···················· | ····················· | ···················· | ···················· | ···················· |
| T | 4096 hCoV-19/Denmark/DCGC-11730/2020\|EPI_ISL_670681\|202 | ···················· | ···················· | ····················· | ···················· | ···················· | ···················· |
| T | 4097 hCoV-19/England/QEUH-B67479/2020\|EPI_ISL_662136\|20 | ···················· | ···················· | ····················· | ···················· | ···················· | ···················· |
| T | 4098 hCoV-19/England/ALDP-B744A6/2020\|EPI_ISL_662137\|20 | ···················· | ···················· | ····················· | ···················· | ···················· | ···················· |
| T | 4099 hCoV-19/England/ALDP-B74FE1/2020\|EPI_ISL_662138\|20 | ···················· | ···················· | ····················· | ···················· | ···················· | ···················· |
| T | 4100 hCoV-19/England/QEUH-B68715/2020\|EPI_ISL_662139\|20 | ···················· | ···················· | ····················· | ···················· | ···················· | ···················· |
| T | 4101 hCoV-19/England/ALDP-B74682/2020\|EPI_ISL_662140\|20 | ···················· | ···················· | ····················· | ···················· | ···················· | ···················· |
| T | 4102 hCoV-19/England/ALDP-B75056/2020\|EPI_ISL_662141\|20 | ···················· | ···················· | ····················· | ···················· | ···················· | ···················· |
| T | 4103 hCoV-19/England/QEUH-B66E9A/2020\|EPI_ISL_662142\|20 | ···················· | ···················· | ····················· | ···················· | ···················· | ···················· |
| T | 4104 hCoV-19/England/ALDP-B751BD/2020\|EPI_ISL_662143\|20 | ···················· | ···················· | ····················· | ···················· | ···················· | ···················· |
| T | 4105 hCoV-19/England/QEUH-B673F4/2020\|EPI_ISL_662144\|20 | ···················· | ···················· | ····················· | ···················· | ···················· | ···················· |
| T | 4106 hCoV-19/England/ALDP-B7519F/2020\|EPI_ISL_662145\|20 | ···················· | ···················· | ····················· | ···················· | ···················· | ···················· |
| T | 4107 hCoV-19/England/ALDP-B73FA6/2020\|EPI_ISL_662146\|20 | ···················· | ···················· | ····················· | ···················· | ···················· | ···················· |
| T | 4108 hCoV-19/England/QEUH-B67F69/2020\|EPI_ISL_662147\|20 | ···················· | ···················· | ····················· | ···················· | ···················· | ···················· |
| T | 4109 hCoV-19/England/ALDP-B74CF9/2020\|EPI_ISL_662148\|20 | ···················· | ···················· | ····················· | ···················· | ···················· | ···················· |
| T | 4110 hCoV-19/England/QEUH-B686DC/2020\|EPI_ISL_662149\|20 | ···················· | ···················· | ····················· | ···················· | ···················· | ···················· |
| T | 4111 hCoV-19/England/ALDP-B7436D/2020\|EPI_ISL_662150\|20 | ···················· | ···················· | ····················· | ···················· | ···················· | ···················· |
| T | 4112 hCoV-19/England/ALDP-B73FF1/2020\|EPI_ISL_662151\|20 | ···················· | ···················· | ····················· | ···················· | ···················· | ···················· |
| T | 4113 hCoV-19/England/QEUH-B68979/2020\|EPI_ISL_662152\|20 | ···················· | ···················· | ····················· | ···················· | ···················· | ···················· |
| T | 4114 hCoV-19/England/QEUH-B6891F/2020\|EPI_ISL_662153\|20 | ···················· | ···················· | ····················· | ···················· | ···················· | ···················· |
| T | 4115 hCoV-19/England/QEUH-B680CF/2020\|EPI_ISL_662154\|20 | ···················· | ···················· | ····················· | ···················· | ···················· | ···················· |
| T | 4116 hCoV-19/England/QEUH-B685FD/2020\|EPI_ISL_662155\|20 | ···················· | ···················· | ····················· | ···················· | ···················· | ···················· |

| | | | | | | | | |
|---|---|---|---|---|---|---|---|---|
| T | 4117 | hCoV-19/England/ALDP-B74752/2020\|EPI_ISL_662156\|20 | ···················· | ···················· | ···················· | ···················· | ···················· | ···················· |
| T | 4118 | hCoV-19/England/QEUH-B68724/2020\|EPI_ISL_662157\|20 | ···················· | ···················· | ···················· | ···················· | ···················· | ···················· |
| T | 4119 | hCoV-19/England/QEUH-B69204/2020\|EPI_ISL_662158\|20 | ···················· | ···················· | ···················· | ···················· | ···················· | ···················· |
| T | 4120 | hCoV-19/England/ALDP-B749A7/2020\|EPI_ISL_662159\|20 | ···················· | ···················· | ···················· | ···················· | ···················· | ···················· |
| T | 4121 | hCoV-19/England/QEUH-B69134/2020\|EPI_ISL_662161\|20 | ···················· | ···················· | ···················· | ···················· | ···················· | ···················· |
| T | 4122 | hCoV-19/England/ALDP-B75609/2020\|EPI_ISL_662162\|20 | ···················· | ···················· | ···················· | ···················· | ···················· | ···················· |
| T | 4123 | hCoV-19/England/ALDP-B74E6C/2020\|EPI_ISL_662163\|20 | ···················· | ···················· | ···················· | ···················· | ···················· | ···················· |
| T | 4124 | hCoV-19/England/ALDP-B74497/2020\|EPI_ISL_662164\|20 | ···················· | ···················· | ···················· | ···················· | ···················· | ···················· |
| T | 4125 | hCoV-19/England/ALDP-B746A0/2020\|EPI_ISL_662165\|20 | ···················· | ···················· | ···················· | ···················· | ···················· | ···················· |
| T | 4126 | hCoV-19/England/ALDP-B74637/2020\|EPI_ISL_662166\|20 | ···················· | ···················· | ···················· | ···················· | ···················· | ···················· |
| T | 4127 | hCoV-19/England/ALDP-B752F6/2020\|EPI_ISL_662167\|20 | ···················· | ···················· | ···················· | ···················· | ···················· | ···················· |
| T | 4128 | hCoV-19/England/QEUH-B68162/2020\|EPI_ISL_662168\|20 | ···················· | ···················· | ···················· | ···················· | ···················· | ···················· |
| T | 4129 | hCoV-19/England/ALDP-B74D14/2020\|EPI_ISL_662169\|20 | ···················· | ···················· | ···················· | ···················· | ···················· | ···················· |
| T | 4130 | hCoV-19/England/ALDP-B74CBD/2020\|EPI_ISL_662170\|20 | ···················· | ···················· | ···················· | ···················· | ···················· | ···················· |
| T | 4131 | hCoV-19/England/QEUH-B68EC5/2020\|EPI_ISL_662171\|20 | ···················· | ···················· | ···················· | ···················· | ···················· | ···················· |
| T | 4132 | hCoV-19/England/QEUH-B68F68/2020\|EPI_ISL_662172\|20 | ···················· | ···················· | ···················· | ···················· | ···················· | ···················· |
| T | 4133 | hCoV-19/England/ALDP-B7536C/2020\|EPI_ISL_662174\|20 | ···················· | ···················· | ···················· | ···················· | ···················· | ···················· |
| T | 4134 | hCoV-19/England/ALDP-B751CC/2020\|EPI_ISL_662175\|20 | ···················· | ···················· | ···················· | ···················· | ···················· | ···················· |
| T | 4135 | hCoV-19/England/ALDP-B746BF/2020\|EPI_ISL_662176\|20 | ···················· | ···················· | ···················· | ···················· | ···················· | ···················· |
| T | 4136 | hCoV-19/England/ALDP-B75320/2020\|EPI_ISL_662177\|20 | ···················· | ···················· | ···················· | ···················· | ···················· | ···················· |
| T | 4137 | hCoV-19/England/ALDP-B75153/2020\|EPI_ISL_662178\|20 | ···················· | ···················· | ···················· | ···················· | ···················· | ···················· |
| T | 4138 | hCoV-19/England/ALDP-B73F97/2020\|EPI_ISL_662179\|20 | ···················· | ···················· | ···················· | ···················· | ···················· | ···················· |
| T | 4139 | hCoV-19/England/QEUH-B691CB/2020\|EPI_ISL_662180\|20 | ···················· | ···················· | ···················· | ···················· | ···················· | ···················· |
| T | 4140 | hCoV-19/England/ALDP-B74CEA/2020\|EPI_ISL_662181\|20 | ···················· | ···················· | ···················· | ···················· | ···················· | ···················· |
| T | 4141 | hCoV-19/England/ALDP-B75645/2020\|EPI_ISL_662183\|20 | ···················· | ···················· | ···················· | ···················· | ···················· | ···················· |
| T | 4142 | hCoV-19/England/ALDP-B744D3/2020\|EPI_ISL_662184\|20 | ···················· | ···················· | ···················· | ···················· | ···················· | ···················· |
| T | 4143 | hCoV-19/England/ALDP-B75092/2020\|EPI_ISL_662186\|20 | ···················· | ···················· | ···················· | ···················· | ···················· | ···················· |
| T | 4144 | hCoV-19/England/ALDP-B744E2/2020\|EPI_ISL_662187\|20 | ···················· | ···················· | ···················· | ···················· | ···················· | ···················· |
| T | 4145 | hCoV-19/England/QEUH-B6801A/2020\|EPI_ISL_662188\|20 | ···················· | ·A·················· | ···················· | ···················· | ···················· | ···················· |
| T | 4146 | hCoV-19/England/QEUH-B674D3/2020\|EPI_ISL_662189\|20 | ···················· | ···················· | ···················· | ···················· | ···················· | ···················· |
| T | 4147 | hCoV-19/England/QEUH-B6739A/2020\|EPI_ISL_662190\|20 | ···················· | ···················· | ···················· | ···················· | ···················· | ···················· |
| T | 4148 | hCoV-19/England/ALDP-B74761/2020\|EPI_ISL_662191\|20 | ···················· | ···················· | ···················· | ···················· | ···················· | ···················· |
| T | 4149 | hCoV-19/England/ALDP-B7533F/2020\|EPI_ISL_662192\|20 | ···················· | ···················· | ···················· | ···················· | ···················· | ···················· |
| T | 4150 | hCoV-19/England/ALDP-B75733/2020\|EPI_ISL_662194\|20 | ···················· | ·A·················· | ···················· | ···················· | ···················· | ···················· |
| T | 4151 | hCoV-19/England/QEUH-B68029/2020\|EPI_ISL_662195\|20 | ···················· | ·A·················· | ···················· | ···················· | ···················· | ···················· |
| T | 4152 | hCoV-19/England/ALDP-B7579D/2020\|EPI_ISL_662196\|20 | ···················· | ·A·················· | ···················· | ···················· | ···················· | ···················· |
| T | 4153 | hCoV-19/England/QEUH-B686BE/2020\|EPI_ISL_662197\|20 | ···················· | ···················· | ···················· | ···················· | ···················· | ···················· |
| T | 4154 | hCoV-19/England/ALDP-B74084/2020\|EPI_ISL_662198\|20 | ···················· | ···················· | ···················· | ···················· | ···················· | ···················· |
| T | 4155 | hCoV-19/England/QEUH-B68D40/2020\|EPI_ISL_662199\|20 | ···················· | ···················· | ···················· | ···················· | ···················· | ···················· |
| T | 4156 | hCoV-19/England/ALDP-B746DD/2020\|EPI_ISL_662200\|20 | ···················· | ···················· | ···················· | ···················· | ···················· | ···················· |
| T | 4157 | hCoV-19/England/QEUH-B69143/2020\|EPI_ISL_662201\|20 | ···················· | ···················· | ···················· | ···················· | ···················· | ···················· |
| T | 4158 | hCoV-19/England/ALDP-B755FD/2020\|EPI_ISL_662202\|20 | ···················· | ···················· | ···················· | ···················· | ···················· | ···················· |
| T | 4159 | hCoV-19/England/QEUH-B69161/2020\|EPI_ISL_662204\|20 | ···················· | ·A·················· | ···················· | ···················· | ···················· | ··A················· |
| T | 4160 | hCoV-19/England/QEUH-B680B0/2020\|EPI_ISL_662205\|20 | ···················· | ···················· | ···················· | ···················· | ···················· | ···················· |
| T | 4161 | hCoV-19/England/ALDP-B75706/2020\|EPI_ISL_662206\|20 | ···················· | ···················· | ···················· | ···················· | ···················· | ···················· |
| T | 4162 | hCoV-19/England/ALDP-B7400C/2020\|EPI_ISL_662208\|20 | ···················· | ···················· | ···················· | ···················· | ···················· | ···················· |
| T | 4163 | hCoV-19/England/ALDP-B75135/2020\|EPI_ISL_662209\|20 | ···················· | ···················· | ···················· | ···················· | ···················· | ···················· |
| T | 4164 | hCoV-19/England/QEUH-B68F77/2020\|EPI_ISL_662210\|20 | ···················· | ···················· | ···················· | ···················· | ···················· | T··················· |
| T | 4165 | hCoV-19/England/QEUH-B68FA4/2020\|EPI_ISL_662211\|20 | ···················· | ···················· | ···················· | ···················· | ···················· | ···················· |
| T | 4166 | hCoV-19/England/ALDP-B756EB/2020\|EPI_ISL_662212\|20 | ···················· | ···················· | ···················· | ···················· | ···················· | ···················· |
| T | 4167 | hCoV-19/England/QEUH-B686AF/2020\|EPI_ISL_662222\|20 | ···················· | ···················· | ···················· | ···················· | ···················· | ···················· |
| T | 4168 | hCoV-19/England/ALDP-B73FB5/2020\|EPI_ISL_662223\|20 | ···················· | ···················· | ···················· | ···················· | ···················· | ···················· |
| T | 4169 | hCoV-19/Australia/VIC16905/2020\|EPI_ISL_663529\|202 | ···················· | ···················· | ···················· | ···················· | ···················· | ···················· |
| T | 4172 | hCoV-19/Australia/VIC16924/2020\|EPI_ISL_663532\|202 | ···················· | ···················· | ···················· | ···················· | ···················· | ···················· |
| T | 4173 | hCoV-19/Australia/VIC16925/2020\|EPI_ISL_663533\|202 | ···················· | ···················· | ···················· | ···················· | ···················· | ···················· |
| T | 4174 | hCoV-19/Australia/VIC16927/2020\|EPI_ISL_663534\|202 | ···················· | ···················· | ···················· | ···················· | ···················· | ···················· |
| T | 4176 | hCoV-19/Australia/VIC16904/2020\|EPI_ISL_663536\|202 | ···················· | ···················· | ···················· | ···················· | NN·················· | ···················· |
| T | 4178 | hCoV-19/Australia/VIC16988/2020\|EPI_ISL_663538\|202 | ···················· | ···················· | ···················· | ···················· | ···················· | ···················· |
| T | 4180 | hCoV-19/Australia/VIC17361/2020\|EPI_ISL_663540\|202 | ···················· | ···················· | ···················· | ···················· | ···················· | ···················· |
| T | 4182 | hCoV-19/Australia/VIC17501/2020\|EPI_ISL_663542\|202 | ···················· | ···················· | ···················· | ···················· | ···················· | ···················· |
| T | 4183 | hCoV-19/Australia/VIC17567/2020\|EPI_ISL_663543\|202 | ···················· | ···················· | ···················· | ···················· | ···················· | ···················· |
| T | 4185 | hCoV-19/Australia/VIC16728/2020\|EPI_ISL_663554\|202 | ···················· | ···················· | ···················· | ···················· | ···················· | ···················· |
| T | 4186 | hCoV-19/England/ALDP-B1CA35/2020\|EPI_ISL_664840\|20 | ···················· | ···················· | ···················· | ···················· | ···················· | ···················· |
| T | 4187 | hCoV-19/England/ALDP-B1CA44/2020\|EPI_ISL_664841\|20 | ···················· | ···················· | ···················· | ···················· | ···················· | ···················· |
| T | 4188 | hCoV-19/England/ALDP-B1CA62/2020\|EPI_ISL_664842\|20 | ···················· | ···················· | ···················· | ···················· | ·····T·············· | ···················· |
| T | 4189 | hCoV-19/England/ALDP-B1CA71/2020\|EPI_ISL_664843\|20 | ···················· | ···················· | ···················· | ···················· | ···················· | ···················· |
| T | 4190 | hCoV-19/England/ALDP-B1CA80/2020\|EPI_ISL_664844\|20 | ···················· | ···················· | ···················· | ···················· | ···················· | ···················· |
| T | 4191 | hCoV-19/England/ALDP-B1CADB/2020\|EPI_ISL_664845\|20 | ···················· | ···················· | ···················· | ···················· | ···················· | ···················· |
| T | 4192 | hCoV-19/England/ALDP-B1CB41/2020\|EPI_ISL_664846\|20 | ···················· | ···················· | ···················· | ···················· | ···················· | ···················· |
| T | 4193 | hCoV-19/England/ALDP-B1CBBA/2020\|EPI_ISL_664847\|20 | ···················· | ···················· | ···················· | ···················· | ···················· | ···················· |
| T | 4194 | hCoV-19/England/ALDP-B1CBF6/2020\|EPI_ISL_664848\|20 | ···················· | ···················· | ···················· | ···················· | ···················· | ···················· |
| T | 4195 | hCoV-19/England/ALDP-B1CC4E/2020\|EPI_ISL_664849\|20 | ···················· | ···················· | ···················· | ···················· | ···················· | ···················· |
| T | 4196 | hCoV-19/England/ALDP-B1CC5D/2020\|EPI_ISL_664850\|20 | ···················· | ···················· | ···················· | ···················· | ···················· | ···················· |
| T | 4197 | hCoV-19/England/ALDP-B1D023/2020\|EPI_ISL_664851\|20 | ···················· | ···················· | ···················· | ···················· | ···················· | ···················· |
| T | 4198 | hCoV-19/England/ALDP-B1D07E/2020\|EPI_ISL_664852\|20 | ···················· | ···················· | ···················· | ···················· | ···················· | ···················· |
| T | 4199 | hCoV-19/England/ALDP-B1D08D/2020\|EPI_ISL_664853\|20 | ···················· | ···················· | ···················· | ···················· | ···················· | ···················· |
| T | 4200 | hCoV-19/England/ALDP-B1D0C9/2020\|EPI_ISL_664854\|20 | ···················· | ···················· | ···················· | ···················· | ···················· | ···················· |
| T | 4201 | hCoV-19/England/ALDP-B1D111/2020\|EPI_ISL_664855\|20 | ···················· | ···················· | ···················· | ···················· | ···················· | ···················· |
| T | 4202 | hCoV-19/England/ALDP-B271AB/2020\|EPI_ISL_664856\|20 | ···················· | ···················· | ···················· | ···················· | ···················· | ···················· |
| T | 4203 | hCoV-19/England/ALDP-B737AE/2020\|EPI_ISL_664857\|20 | ···················· | ···················· | ···················· | ···················· | ···················· | ···················· |

| | | | | | | | | |
|---|---|---|---|---|---|---|---|---|
| T | 4204 hCoV-19/England/ALDP-B737CC/2020\|EPI_ISL_664858\|20 | ···················· | ···················· | ···················· | ···················· | ···················· | ···················· | |
| T | 4205 hCoV-19/England/ALDP-B737EA/2020\|EPI_ISL_664859\|20 | ···················· | ···················· | ···················· | ···················· | ···················· | ···················· | |
| T | 4206 hCoV-19/England/ALDP-B737F9/2020\|EPI_ISL_664860\|20 | ···················· | ···················· | ···················· | ···················· | ···················· | ···················· | |
| T | 4207 hCoV-19/England/ALDP-B73823/2020\|EPI_ISL_664861\|20 | ···················· | ···················· | ···················· | ···················· | ···················· | ···················· | |
| T | 4208 hCoV-19/England/ALDP-B73832/2020\|EPI_ISL_664862\|20 | ···················· | ···················· | ···················· | ···················· | N··················· | ···················· | |
| T | 4210 hCoV-19/England/ALDP-B73999/2020\|EPI_ISL_664864\|20 | ···················· | ···················· | ···················· | ···················· | ···················· | ···················· | |
| T | 4213 hCoV-19/England/ALDP-B739E4/2020\|EPI_ISL_664867\|20 | ···················· | ···················· | ···················· | ···················· | N··················· | ···················· | |
| T | 4214 hCoV-19/England/ALDP-B73A96/2020\|EPI_ISL_664868\|20 | ···················· | ···················· | ···················· | ···················· | ···················· | ···················· | |
| T | 4216 hCoV-19/England/ALDP-B73AF0/2020\|EPI_ISL_664870\|20 | ···················· | ···················· | ···················· | ···················· | ···················· | ···················· | |
| T | 4218 hCoV-19/England/ALDP-B73BFD/2020\|EPI_ISL_664872\|20 | ···················· | ···················· | ···················· | ···················· | NNN················· | NNN················· | 1 |
| T | 4219 hCoV-19/England/ALDP-B73C18/2020\|EPI_ISL_664873\|20 | ···················· | ···················· | ···················· | ···················· | ···················· | ···················· | |
| T | 4221 hCoV-19/England/ALDP-B73D24/2020\|EPI_ISL_664884\|20 | ···················· | ···················· | ···················· | ···················· | ···················· | ···················· | |
| T | 4222 hCoV-19/USA/CA-ALSR-4543/2020\|EPI_ISL_667001\|2020- | ···················· | ···················· | ···················· | ···················· | ···················· | ···················· | |
| T | 4223 hCoV-19/USA/CA-ALSR-4564/2020\|EPI_ISL_667009\|2020- | ···················· | ·A·················· | ···················· | ···················· | ···················· | ···················· | |
| T | 4224 hCoV-19/USA/CA-ALSR-4565/2020\|EPI_ISL_667010\|2020- | ···················· | ·A·················· | ···················· | ···················· | ···················· | ···················· | |
| T | 4225 hCoV-19/USA/CA-ALSR-4567/2020\|EPI_ISL_667011\|2020- | ···················· | ·A·················· | ···················· | ···················· | ···················· | ···················· | |
| T | 4226 hCoV-19/USA/CA-ALSR-4570/2020\|EPI_ISL_667012\|2020- | ···················· | ···················· | ···················· | ···················· | ···················· | ···················· | |
| T | 4227 hCoV-19/USA/CA-ALSR-4571/2020\|EPI_ISL_667013\|2020- | ···················· | ·A·················· | ···················· | ···················· | ···················· | ···················· | |
| T | 4228 hCoV-19/USA/CA-ALSR-4574/2020\|EPI_ISL_667014\|2020- | ···················· | ·A·················· | ···················· | ···················· | ···················· | ···················· | |
| T | 4229 hCoV-19/USA/CA-ALSR-4575/2020\|EPI_ISL_667015\|2020- | ···················· | ·A·················· | ···················· | ···················· | ···················· | ···················· | |
| T | 4230 hCoV-19/USA/CA-ALSR-4577/2020\|EPI_ISL_667017\|2020- | ···················· | ···················· | ···················· | ···················· | ···················· | ···················· | |
| T | 4231 hCoV-19/USA/CA-ALSR-4583/2020\|EPI_ISL_667020\|2020- | ···················· | ·A·················· | ···················· | ···················· | ···················· | ···················· | |
| T | 4232 hCoV-19/USA/CA-ALSR-4585/2020\|EPI_ISL_667021\|2020- | ···················· | ·A·················· | ···················· | ···················· | ···················· | ···················· | |
| T | 4233 hCoV-19/USA/CA-ALSR-4586/2020\|EPI_ISL_667022\|2020- | ···················· | ···················· | ···················· | ···················· | ···················· | ···················· | |
| T | 4234 hCoV-19/USA/CA-ALSR-4587/2020\|EPI_ISL_667023\|2020- | ···················· | ·A·················· | ···················· | ···················· | ···················· | ···················· | |
| T | 4235 hCoV-19/USA/CA-ALSR-4588/2020\|EPI_ISL_667024\|2020- | ···················· | ·A·················· | ···················· | ···················· | ···················· | ···················· | |
| T | 4236 hCoV-19/USA/CA-ALSR-4590/2020\|EPI_ISL_667025\|2020- | ···················· | ···················· | ···················· | ···················· | ···················· | ···················· | |
| T | 4237 hCoV-19/USA/CA-ALSR-4593/2020\|EPI_ISL_667027\|2020- | ···················· | ···················· | ···················· | ···················· | ···················· | ···················· | |
| T | 4238 hCoV-19/USA/CA-ALSR-4594/2020\|EPI_ISL_667028\|2020- | ···················· | ···················· | ···················· | ···················· | ···················· | ···················· | |
| T | 4239 hCoV-19/USA/CA-ALSR-4596/2020\|EPI_ISL_667029\|2020- | ···················· | ···················· | ···················· | ···················· | ···················· | ···················· | |
| T | 4240 hCoV-19/England/CAMC-B77661/2020\|EPI_ISL_668329\|20 | ···················· | ···················· | ···················· | ···················· | ···················· | ···················· | |
| T | 4241 hCoV-19/England/QEUH-B7E8A0/2020\|EPI_ISL_668330\|20 | ···················· | ···················· | ···················· | ···················· | ···················· | ···················· | |
| T | 4242 hCoV-19/England/QEUH-B7F933/2020\|EPI_ISL_668331\|20 | ···················· | ···················· | ···················· | ···················· | ···················· | ···················· | |
| T | 4243 hCoV-19/England/QEUH-B8219F/2020\|EPI_ISL_668332\|20 | ···················· | ···················· | ···················· | ···················· | ···················· | ···················· | |
| T | 4244 hCoV-19/England/QEUH-B81400/2020\|EPI_ISL_668333\|20 | ···················· | ···················· | ···················· | ···················· | ···················· | ···················· | |
| T | 4245 hCoV-19/England/QEUH-B7C0B9/2020\|EPI_ISL_668334\|20 | ···················· | ···················· | ···················· | ···················· | ···················· | ···················· | |
| T | 4246 hCoV-19/England/QEUH-B7C198/2020\|EPI_ISL_668340\|20 | ···················· | ···················· | ···················· | ···················· | ···················· | ···················· | |
| T | 4247 hCoV-19/England/QEUH-B7C329/2020\|EPI_ISL_668344\|20 | ···················· | ···················· | ···················· | ···················· | ···················· | ···················· | |
| T | 4248 hCoV-19/England/CAMC-B77FA2/2020\|EPI_ISL_668345\|20 | ···················· | ···················· | ···················· | ···················· | ···················· | ···················· | |
| T | 4249 hCoV-19/England/QEUH-B7E837/2020\|EPI_ISL_668346\|20 | ···················· | ···················· | ···················· | ···················· | ···················· | ···················· | |
| T | 4250 hCoV-19/England/QEUH-B7C20E/2020\|EPI_ISL_668348\|20 | ···················· | ···················· | ···················· | ···················· | ···················· | ···················· | |
| T | 4251 hCoV-19/England/QEUH-B7F65A/2020\|EPI_ISL_668350\|20 | ···················· | ···················· | ···················· | ···················· | ···················· | ···················· | |
| T | 4252 hCoV-19/England/QEUH-B821DB/2020\|EPI_ISL_668352\|20 | ···················· | ···················· | ···················· | ···················· | ···················· | ···················· | |
| T | 4253 hCoV-19/England/CAMC-B77FDF/2020\|EPI_ISL_668353\|20 | ···················· | ···················· | ···················· | ···················· | ···················· | ···················· | |
| T | 4254 hCoV-19/England/CAMC-B77652/2020\|EPI_ISL_668355\|20 | ···················· | ···················· | ···················· | ···················· | ···················· | ···················· | |
| T | 4255 hCoV-19/England/QEUH-B7BFEA/2020\|EPI_ISL_668359\|20 | ···················· | ···················· | ···················· | ···················· | ···················· | ···················· | |
| T | 4256 hCoV-19/Denmark/DCGC-11130/2020\|EPI_ISL_669600\|202 | ···················· | ·A·················· | ···················· | ···················· | ···················· | ···················· | |
| T | 4257 hCoV-19/Denmark/DCGC-10414/2020\|EPI_ISL_669660\|202 | ···················· | ···················· | ···················· | ···················· | ···················· | ···················· | |
| T | 4258 hCoV-19/Denmark/DCGC-10419/2020\|EPI_ISL_669661\|202 | ···················· | ···················· | ···················· | ···················· | ···················· | ···················· | |
| T | 4259 hCoV-19/Denmark/DCGC-10421/2020\|EPI_ISL_669662\|202 | ···················· | ···················· | ···················· | ···················· | ···················· | ···················· | |
| T | 4260 hCoV-19/Denmark/DCGC-10424/2020\|EPI_ISL_669663\|202 | ···················· | ···················· | ···················· | ···················· | ···················· | ···················· | |
| T | 4261 hCoV-19/Denmark/DCGC-10430/2020\|EPI_ISL_669664\|202 | ···················· | ···················· | ···················· | ···················· | ···················· | ···················· | |
| T | 4262 hCoV-19/Denmark/DCGC-10433/2020\|EPI_ISL_669665\|202 | ···················· | ···················· | ···················· | ···················· | ···················· | ···················· | |
| T | 4263 hCoV-19/Denmark/DCGC-10450/2020\|EPI_ISL_669666\|202 | ···················· | ···················· | ···················· | ···················· | ···················· | ···················· | |
| T | 4264 hCoV-19/Denmark/DCGC-10501/2020\|EPI_ISL_669670\|202 | ···················· | ·A·················· | ···················· | ···················· | ···················· | ···················· | |
| T | 4265 hCoV-19/Denmark/DCGC-10507/2020\|EPI_ISL_669671\|202 | ···················· | ···················· | ···················· | ···················· | ···················· | ···················· | |
| T | 4266 hCoV-19/Denmark/DCGC-10516/2020\|EPI_ISL_669672\|202 | ···················· | ···················· | ···················· | ···················· | ···················· | ···················· | |
| T | 4267 hCoV-19/Denmark/DCGC-10527/2020\|EPI_ISL_669673\|202 | ···················· | ···················· | ···················· | ···················· | ···················· | ···················· | |
| T | 4268 hCoV-19/Denmark/DCGC-10529/2020\|EPI_ISL_669674\|202 | ···················· | ···················· | ···················· | ···················· | ···················· | ···················· | |
| T | 4269 hCoV-19/Denmark/DCGC-10532/2020\|EPI_ISL_669675\|202 | ···················· | ···················· | ···················· | ···················· | ···················· | ···················· | |
| T | 4270 hCoV-19/Denmark/DCGC-10534/2020\|EPI_ISL_669676\|202 | ···················· | ···················· | ···················· | ···················· | ···················· | ···················· | |
| T | 4271 hCoV-19/Denmark/DCGC-10582/2020\|EPI_ISL_669677\|202 | ···················· | ···················· | ···················· | ···················· | ···················· | ···················· | |
| T | 4272 hCoV-19/Denmark/DCGC-10593/2020\|EPI_ISL_669678\|202 | ···················· | ···················· | ···················· | ···················· | ···················· | ···················· | |
| T | 4273 hCoV-19/Denmark/DCGC-10606/2020\|EPI_ISL_669679\|202 | ···················· | ···················· | ···················· | ···················· | ···················· | ···················· | |
| T | 4274 hCoV-19/Denmark/DCGC-10630/2020\|EPI_ISL_669680\|202 | ···················· | ···················· | ···················· | ···················· | ···················· | ···················· | |
| T | 4275 hCoV-19/Denmark/DCGC-10632/2020\|EPI_ISL_669681\|202 | ···················· | ···················· | ···················· | ···················· | ···················· | ···················· | |
| T | 4276 hCoV-19/Denmark/DCGC-10655/2020\|EPI_ISL_669682\|202 | ···················· | ···················· | ···················· | ···················· | ···················· | ···················· | |
| T | 4277 hCoV-19/Denmark/DCGC-10660/2020\|EPI_ISL_669683\|202 | ···················· | ···················· | ···················· | ···················· | ···················· | ···················· | |
| T | 4278 hCoV-19/Denmark/DCGC-10667/2020\|EPI_ISL_669684\|202 | ···················· | ···················· | ···················· | ···················· | ···················· | ···················· | |
| T | 4279 hCoV-19/Denmark/DCGC-10688/2020\|EPI_ISL_669685\|202 | ···················· | ···················· | ···················· | ···················· | ···················· | ···················· | |
| T | 4280 hCoV-19/Denmark/DCGC-10724/2020\|EPI_ISL_669686\|202 | ···················· | ···················· | ···················· | ···················· | ···················· | ···················· | |
| T | 4281 hCoV-19/Denmark/DCGC-10741/2020\|EPI_ISL_669687\|202 | ···················· | ···················· | ···················· | ···················· | ···················· | ···················· | |
| T | 4282 hCoV-19/Denmark/DCGC-10757/2020\|EPI_ISL_669688\|202 | ···················· | ···················· | ···················· | ···················· | ···················· | ···················· | |
| T | 4283 hCoV-19/Denmark/DCGC-10758/2020\|EPI_ISL_669689\|202 | ···················· | ···················· | ···················· | ···················· | ···················· | ···················· | |
| T | 4284 hCoV-19/Denmark/DCGC-11848/2020\|EPI_ISL_670640\|202 | ···················· | ···················· | ···················· | ···················· | ···················· | ···················· | |
| T | 4285 hCoV-19/Denmark/DCGC-11879/2020\|EPI_ISL_670641\|202 | ···················· | ···················· | ···················· | ···················· | ···················· | ···················· | |
| T | 4286 hCoV-19/Denmark/DCGC-11886/2020\|EPI_ISL_670642\|202 | ···················· | ···················· | ···················· | ···················· | ···················· | ···················· | |
| T | 4287 hCoV-19/Denmark/DCGC-11888/2020\|EPI_ISL_670643\|202 | ···················· | ···················· | ···················· | ···················· | ···················· | ···················· | |
| T | 4288 hCoV-19/Denmark/DCGC-11920/2020\|EPI_ISL_670644\|202 | ···················· | ···················· | ···················· | ···················· | ···················· | ···················· | |
| T | 4289 hCoV-19/Denmark/DCGC-11925/2020\|EPI_ISL_670645\|202 | ···················· | ···················· | ···················· | ···················· | ···················· | ···················· | |

| | | | | | | | |
|---|---|---|---|---|---|---|---|
| T | 4290 hCoV-19/Denmark/DCGC-11928/2020\|EPI_ISL_670646\|202 | ···················· | ···················· | ···················· | ···················· | ···················· | ···················· |
| T | 4291 hCoV-19/Denmark/DCGC-11933/2020\|EPI_ISL_670647\|202 | ···················· | ···················· | ···················· | ···················· | ···················· | ···················· |
| T | 4292 hCoV-19/Denmark/DCGC-11944/2020\|EPI_ISL_670648\|202 | ···················· | ···················· | ···················· | ···················· | ···················· | ···················· |
| T | 4293 hCoV-19/Denmark/DCGC-11952/2020\|EPI_ISL_670649\|202 | ···················· | ···················· | ···················· | ···················· | ···················· | ···················· |
| T | 4294 hCoV-19/Denmark/DCGC-11957/2020\|EPI_ISL_670650\|202 | ···················· | ···················· | ···················· | ···················· | ···················· | ···················· |
| T | 4295 hCoV-19/Denmark/DCGC-11960/2020\|EPI_ISL_670651\|202 | ···················· | ···················· | ···················· | ···················· | ···················· | ···················· |
| T | 4296 hCoV-19/Denmark/DCGC-11961/2020\|EPI_ISL_670652\|202 | ···················· | ···················· | ···················· | ···················· | ···················· | ···················· |
| T | 4297 hCoV-19/Denmark/DCGC-11963/2020\|EPI_ISL_670653\|202 | ···················· | ···················· | ···················· | ···················· | ···················· | ···················· |
| T | 4298 hCoV-19/Denmark/DCGC-11968/2020\|EPI_ISL_670654\|202 | ···················· | ···················· | ···················· | ···················· | ···················· | ···················· |
| T | 4299 hCoV-19/Denmark/DCGC-11969/2020\|EPI_ISL_670655\|202 | ···················· | ···················· | ···················· | ···················· | ···················· | ···················· |
| T | 4300 hCoV-19/Denmark/DCGC-11970/2020\|EPI_ISL_670656\|202 | ···················· | ···················· | ···················· | ···················· | ···················· | ···················· |
| T | 4301 hCoV-19/Denmark/DCGC-11971/2020\|EPI_ISL_670657\|202 | ···················· | ···················· | ···················· | ···················· | ···················· | ···················· |
| T | 4302 hCoV-19/Denmark/DCGC-11972/2020\|EPI_ISL_670658\|202 | ···················· | ···················· | ···················· | ···················· | ···················· | ···················· |
| T | 4303 hCoV-19/Denmark/DCGC-11974/2020\|EPI_ISL_670659\|202 | ···················· | ···················· | ···················· | ···················· | ···················· | ···················· |
| T | 4304 hCoV-19/Denmark/DCGC-11975/2020\|EPI_ISL_670660\|202 | ···················· | ···················· | ···················· | ···················· | ···················· | ···················· |
| T | 4305 hCoV-19/Denmark/DCGC-11977/2020\|EPI_ISL_670661\|202 | ···················· | ···················· | ···················· | ···················· | ···················· | ···················· |
| T | 4306 hCoV-19/Denmark/DCGC-11978/2020\|EPI_ISL_670662\|202 | ···················· | ···················· | ···················· | ···················· | ···················· | ···················· |
| T | 4307 hCoV-19/Denmark/DCGC-11980/2020\|EPI_ISL_670663\|202 | ···················· | ···················· | ···················· | ···················· | ···················· | ···················· |
| T | 4308 hCoV-19/Denmark/DCGC-11983/2020\|EPI_ISL_670664\|202 | ···················· | ···················· | ···················· | ···················· | ···················· | ···················· |
| T | 4309 hCoV-19/Denmark/DCGC-11985/2020\|EPI_ISL_670665\|202 | ···················· | ···················· | ···················· | ···················· | ···················· | ···················· |
| T | 4310 hCoV-19/Denmark/DCGC-11246/2020\|EPI_ISL_670667\|202 | ···················· | ···················· | ···················· | ···················· | ···················· | ···················· |
| T | 4311 hCoV-19/Denmark/DCGC-11335/2020\|EPI_ISL_670668\|202 | ···················· | ···················· | ···················· | ···················· | ···················· | ···················· |
| T | 4312 hCoV-19/Denmark/DCGC-11531/2020\|EPI_ISL_670669\|202 | ···················· | ·A·················· | ···················· | ···················· | ···················· | ···················· |
| T | 4313 hCoV-19/Denmark/DCGC-11742/2020\|EPI_ISL_670670\|202 | ···················· | ·A·················· | ···················· | ···················· | ···················· | ··A················· |
| T | 4314 hCoV-19/Denmark/DCGC-11816/2020\|EPI_ISL_670671\|202 | ···················· | ·A·················· | ···················· | ···················· | ···················· | ··A················· |
| T | 4315 hCoV-19/Ireland/D-NVRL-75IRL88794/2020\|EPI_ISL_671 | ···················· | ···················· | ···················· | ···················· | ···················· | ···················· |
| T | 4316 hCoV-19/Ireland/D-NVRL-75IRL90324/2020\|EPI_ISL_671 | ···················· | ···················· | ···················· | ···················· | ···················· | ···················· |
| T | 4317 hCoV-19/Ireland/G-NVRL-75IRL71302/2020\|EPI_ISL_671 | ···················· | ···················· | ···················· | ···················· | ···················· | ···················· |
| T | 4318 hCoV-19/Ireland/G-NVRL-75IRL85497/2020\|EPI_ISL_671 | ···················· | ···················· | ···················· | ···················· | ···················· | ···················· |
| T | 4319 hCoV-19/Ireland/G-NVRL-76IRL13780/2020\|EPI_ISL_671 | ···················· | ···················· | ···················· | ···················· | ···················· | ···················· |
| T | 4320 hCoV-19/Ireland/G-NVRL-76IRL13817/2020\|EPI_ISL_671 | ···················· | ···················· | ···················· | ···················· | ···················· | ···················· |
| T | 4321 hCoV-19/Ireland/G-NVRL-76IRL13842/2020\|EPI_ISL_671 | ···················· | ···················· | ···················· | ···················· | ···················· | ···················· |
| T | 4322 hCoV-19/Ireland/G-NVRL-76IRL13959/2020\|EPI_ISL_671 | ···················· | ···················· | ···················· | ···················· | ···················· | ···················· |
| T | 4323 hCoV-19/Ireland/KE-NVRL-75IRL25866/2020\|EPI_ISL_67 | ···················· | ···················· | ···················· | ···················· | ···················· | ···················· |
| T | 4324 hCoV-19/Ireland/KY-NVRL-75IRL32742/2020\|EPI_ISL_67 | ···················· | ···················· | ···················· | ···················· | ···················· | ···················· |
| T | 4325 hCoV-19/Ireland/KY-NVRL-75IRL93222/2020\|EPI_ISL_67 | ···················· | ···················· | ···················· | ···················· | ···················· | ···················· |
| T | 4326 hCoV-19/Ireland/LH-NVRL-75IRL19162/2020\|EPI_ISL_67 | ···················· | ···················· | ···················· | ···················· | ···················· | ···················· |
| T | 4327 hCoV-19/Ireland/LH-NVRL-75IRL19164/2020\|EPI_ISL_67 | ···················· | ···················· | ···················· | ···················· | ···················· | ···················· |
| T | 4328 hCoV-19/Ireland/LH-NVRL-75IRL19170/2020\|EPI_ISL_67 | ···················· | ···················· | ···················· | ···················· | ···················· | ···················· |
| T | 4329 hCoV-19/Ireland/LK-NVRL-75IRL23364/2020\|EPI_ISL_67 | ···················· | ···················· | ···················· | ···················· | ···················· | ···················· |
| T | 4330 hCoV-19/Ireland/LK-NVRL-75IRL23376/2020\|EPI_ISL_67 | ···················· | ···················· | ···················· | ···················· | ···················· | ···················· |
| T | 4331 hCoV-19/Ireland/LK-NVRL-75IRL23491/2020\|EPI_ISL_67 | ···················· | ···················· | ···················· | ···················· | ···················· | ···················· |
| T | 4332 hCoV-19/Ireland/LK-NVRL-75IRL32062/2020\|EPI_ISL_67 | ···················· | ···················· | ···················· | ···················· | ···················· | ···················· |
| T | 4333 hCoV-19/Ireland/LK-NVRL-75IRL32487/2020\|EPI_ISL_67 | ···················· | ···················· | ···················· | ···················· | ···················· | ···················· |
| T | 4334 hCoV-19/France/OCC-125/2020\|EPI_ISL_671970\|2020-10 | ···················· | ···················· | ···················· | ···················· | ···················· | ···················· |
| T | 4335 hCoV-19/France/OCC-126/2020\|EPI_ISL_671971\|2020-10 | ···················· | ···················· | ···················· | ···················· | ···················· | ···················· |
| T | 4336 hCoV-19/France/OCC-127/2020\|EPI_ISL_671972\|2020-08 | ···················· | ···················· | ···················· | ···················· | ···················· | ···················· |
| T | 4337 hCoV-19/El Salvador/INS-01/2020\|EPI_ISL_671974\|202 | ···················· | ···················· | ···················· | ···················· | ···················· | ···················· |
| T | 4338 hCoV-19/France/OCC-66/2020\|EPI_ISL_671975\|2020-08- | ···················· | ···················· | ···················· | ···················· | ···················· | ···················· |
| T | 4339 hCoV-19/France/OCC-99/2020\|EPI_ISL_671976\|2020-03- | ···················· | ···················· | ···················· | ···················· | ···················· | ···················· |
| T | 4340 hCoV-19/France/OCC-114/2020\|EPI_ISL_671977\|2020-10 | ···················· | ···················· | ···················· | ···················· | ···················· | ···················· |
| T | 4341 hCoV-19/El Salvador/UES-001/2020\|EPI_ISL_671978\|20 | ···················· | ···················· | ···················· | ···················· | ···················· | ···················· |
| T | 4342 hCoV-19/Ecuador/UEES-HE134/2020\|EPI_ISL_671981\|202 | ···················· | ···················· | ···················· | ···················· | ···················· | ···················· |
| T | 4343 hCoV-19/Ecuador/UEES-1921/2020\|EPI_ISL_671983\|2020 | ···················· | ···················· | ···················· | ···················· | ···················· | ···················· |
| T | 4344 hCoV-19/Ecuador/UEES-3911/2020\|EPI_ISL_671984\|2020 | ···················· | ···················· | ···················· | ···················· | ···················· | ···················· |
| T | 4345 hCoV-19/Ecuador/UEES-7352/2020\|EPI_ISL_671985\|2020 | ···················· | ···················· | ···················· | ···················· | ···················· | ···················· |
| T | 4346 hCoV-19/Ecuador/UEES-4040/2020\|EPI_ISL_671988\|2020 | ···················· | ···················· | ···················· | ···················· | ···················· | ···················· |
| T | 4347 hCoV-19/Ecuador/UEES-7435/2020\|EPI_ISL_671990\|2020 | ···················· | ···················· | ···················· | ···················· | ···················· | ···················· |
| T | 4348 hCoV-19/Ecuador/UEES-17/2020\|EPI_ISL_671991\|2020 | ···················· | ···················· | ···················· | ···················· | ···················· | ···················· |
| T | 4349 hCoV-19/Ecuador/UEES-4375/2020\|EPI_ISL_671994\|2020 | ···················· | ···················· | ···················· | ···················· | ···················· | ···················· |
| T | 4350 hCoV-19/Ecuador/UEES-7462/2020\|EPI_ISL_671995\|2020 | ···················· | ···················· | ···················· | ···················· | ···················· | ···················· |
| T | 4351 hCoV-19/Ecuador/UEES-2963/2020\|EPI_ISL_671998\|2020 | ···················· | ···················· | ···················· | ···················· | ···················· | ···················· |
| T | 4352 hCoV-19/Ecuador/UEES-4107/2020\|EPI_ISL_671999\|2020 | ···················· | ···················· | ···················· | ···················· | ···················· | ···················· |
| T | 4353 hCoV-19/England/ALDP-B7534E/2020\|EPI_ISL_662100\|20 | ···················· | ···················· | ···················· | ···················· | ···················· | ···················· |
| T | 4354 hCoV-19/England/ALDP-B74664/2020\|EPI_ISL_662101\|20 | ···················· | ···················· | ···················· | ···················· | ···················· | ···················· |
| T | 4355 hCoV-19/England/QEUH-B67497/2020\|EPI_ISL_662102\|20 | ···················· | ···················· | ···················· | ···················· | ···················· | ···················· |
| T | 4356 hCoV-19/England/QEUH-B67F87/2020\|EPI_ISL_662103\|20 | ···················· | ···················· | ···················· | ···················· | ···················· | ···················· |
| T | 4357 hCoV-19/England/QEUH-B68645/2020\|EPI_ISL_662104\|20 | ···················· | ···················· | ···················· | ···················· | ···················· | ···················· |
| T | 4358 hCoV-19/England/ALDP-B74D8D/2020\|EPI_ISL_662105\|20 | ···················· | ···················· | ···················· | ···················· | ···················· | ···················· |
| T | 4359 hCoV-19/England/ALDP-B755DF/2020\|EPI_ISL_662106\|20 | ···················· | ···················· | ···················· | ···················· | ···················· | ···················· |
| T | 4360 hCoV-19/England/ALDP-B752BA/2020\|EPI_ISL_662107\|20 | ···················· | ···················· | ···················· | ···················· | ···················· | ···················· |
| T | 4361 hCoV-19/England/ALDP-B74707/2020\|EPI_ISL_662108\|20 | ···················· | ···················· | ···················· | ···················· | ···················· | ···················· |
| T | 4362 hCoV-19/England/QEUH-B67F78/2020\|EPI_ISL_662109\|20 | ···················· | ···················· | ···················· | ···················· | ···················· | ···················· |
| T | 4363 hCoV-19/England/QEUH-B680FC/2020\|EPI_ISL_662110\|20 | ···················· | ···················· | ···················· | ···················· | ···················· | ···················· |
| T | 4364 hCoV-19/England/QEUH-B68E89/2020\|EPI_ISL_662111\|20 | ···················· | ···················· | ···················· | ···················· | ···················· | ···················· |
| T | 4365 hCoV-19/England/ALDP-B7535D/2020\|EPI_ISL_662114\|20 | ···················· | ···················· | ···················· | ···················· | ···················· | ···················· |
| T | 4366 hCoV-19/England/ALDP-B75047/2020\|EPI_ISL_662115\|20 | ···················· | ···················· | ···················· | ···················· | ···················· | ···················· |
| T | 4367 hCoV-19/England/ALDP-B74D6F/2020\|EPI_ISL_662116\|20 | ···················· | ···················· | ···················· | ···················· | ···················· | ···················· |
| T | 4368 hCoV-19/England/QEUH-B68609/2020\|EPI_ISL_662117\|20 | ···················· | ···················· | ···················· | ···················· | ······T············· | ···················· |
| T | 4369 hCoV-19/England/ALDP-B74048/2020\|EPI_ISL_662118\|20 | ···················· | ···················· | ···················· | ···················· | ···················· | ···················· |

| | | | | | | | | |
|---|---|---|---|---|---|---|---|---|
| T | 4370 | hCoV-19/England/QEUH-B68D7D/2020\|EPI_ISL_662119\|20 | ···················· | ···················· | ····················· | ···················· | ···················· | ···················· |
| T | 4371 | hCoV-19/England/ALDP-B742AC/2020\|EPI_ISL_662120\|20 | ···················· | ···················· | ····················· | ···················· | ···················· | ···················· |
| T | 4372 | hCoV-19/England/QEUH-B673C7/2020\|EPI_ISL_662121\|20 | ···················· | ···················· | ····················· | ···················· | ···················· | ···················· |
| T | 4373 | hCoV-19/England/ALDP-B7446A/2020\|EPI_ISL_662122\|20 | ···················· | ···················· | ····················· | ···················· | ···················· | ···················· |
| T | 4374 | hCoV-19/England/ALDP-B74C71/2020\|EPI_ISL_662123\|20 | ···················· | ···················· | ····················· | ···················· | ···················· | ···················· |
| T | 4375 | hCoV-19/England/ALDP-B74DD8/2020\|EPI_ISL_662124\|20 | ···················· | ···················· | ····················· | ···················· | ···················· | ···················· |
| T | 4376 | hCoV-19/England/QEUH-B6919E/2020\|EPI_ISL_662125\|20 | ···················· | ···················· | ····················· | ···················· | ···················· | ···················· |
| T | 4377 | hCoV-19/England/QEUH-B66F2E/2020\|EPI_ISL_662126\|20 | ···················· | ···················· | ····················· | ···················· | ···················· | ···················· |
| T | 4378 | hCoV-19/England/QEUH-B68E3E/2020\|EPI_ISL_662127\|20 | ···················· | ···················· | ····················· | ···················· | ···················· | ···················· |
| T | 4379 | hCoV-19/England/QEUH-B66F3D/2020\|EPI_ISL_662128\|20 | ···················· | ···················· | ····················· | ···················· | ···················· | ···················· |
| T | 4380 | hCoV-19/England/ALDP-B755C0/2020\|EPI_ISL_662129\|20 | ···················· | ···················· | ····················· | ···················· | ···················· | ···················· |
| T | 4381 | hCoV-19/England/QEUH-B68EF2/2020\|EPI_ISL_662130\|20 | ···················· | ·A·················· | ····················· | ···················· | ···················· | ···················· |
| T | 4382 | hCoV-19/England/ALDP-B7492F/2020\|EPI_ISL_662131\|20 | ···················· | ···················· | ····················· | ···················· | ···················· | ···················· |
| T | 4383 | hCoV-19/England/ALDP-B7429D/2020\|EPI_ISL_662132\|20 | ···················· | ···················· | ····················· | ···················· | ···················· | ···················· |
| T | 4384 | hCoV-19/England/ALDP-B7479E/2020\|EPI_ISL_662133\|20 | ···················· | ·A·················· | ····················· | ···················· | ···················· | ···············A···· |
| T | 4385 | hCoV-19/England/QEUH-B68DF5/2020\|EPI_ISL_662134\|20 | ···················· | ···················· | ····················· | ···················· | ···················· | ···················· |
| T | 4386 | hCoV-19/England/ALDP-B75144/2020\|EPI_ISL_662135\|20 | ···················· | ···················· | ····················· | ···················· | ···················· | ···················· |
| T | 4387 | hCoV-19/Australia/VIC17211/2020\|EPI_ISL_663437\|202 | ···················· | ···················· | ····················· | ···················· | ···················· | ···················· |
| T | 4388 | hCoV-19/Australia/VIC17219/2020\|EPI_ISL_663438\|202 | ···················· | ···················· | ····················· | ···················· | ···················· | ···················· |
| T | 4389 | hCoV-19/Australia/VIC17606/2020\|EPI_ISL_663439\|202 | ···················· | ···················· | ····················· | ···················· | ···················· | ···················· |
| T | 4390 | hCoV-19/Australia/VIC17309/2020\|EPI_ISL_663440\|202 | ···················· | ···················· | ····················· | ···················· | ···················· | ···················· |
| T | 4392 | hCoV-19/England/LOND-124F99C/2020\|EPI_ISL_664744\|2 | ···················· | ···················· | ····················· | ···················· | ···················· | ···················· |
| T | 4394 | hCoV-19/England/ALDP-B1CC8A/2020\|EPI_ISL_664746\|20 | ···················· | ···················· | ····················· | ···················· | ···················· | ···················· |
| T | 4395 | hCoV-19/England/CAMB-1BA462/2020\|EPI_ISL_664748\|20 | ···················· | ···················· | ····················· | ···················· | ···················· | ···················· |
| T | 4396 | hCoV-19/England/LOND-1250804/2020\|EPI_ISL_664754\|2 | ···················· | ···················· | ····················· | ···················· | ···················· | ···················· |
| T | 4398 | hCoV-19/England/ALDP-B1D06F/2020\|EPI_ISL_664763\|20 | ···················· | ·A·················· | ····················· | ···················· | ···················· | ··A················· |
| T | 4399 | hCoV-19/England/ALDP-B7540F/2020\|EPI_ISL_664764\|20 | ···················· | ·A·················· | ····················· | ···················· | ···················· | ··A················· |
| T | 4400 | hCoV-19/England/CAMB-1BA5F6/2020\|EPI_ISL_664765\|20 | ···················· | ·A·················· | ····················· | ···················· | ···················· | ··A················· |
| T | 4401 | hCoV-19/USA/CA-ALSR-4557/2020\|EPI_ISL_667005\|2020- | ···················· | ·A·················· | ····················· | ···················· | ···················· | ···················· |
| T | 4402 | hCoV-19/USA/CA-ALSR-4561/2020\|EPI_ISL_667007\|2020- | ···················· | ·A·················· | ····················· | ···················· | ···················· | ···················· |
| T | 4403 | hCoV-19/England/QEUH-B7C31A/2020\|EPI_ISL_668240\|20 | ···················· | ···················· | ····················· | ···················· | ···················· | ···················· |
| T | 4404 | hCoV-19/England/QEUH-B7FD37/2020\|EPI_ISL_668241\|20 | ···················· | ···················· | ····················· | ···················· | ···················· | ···················· |
| T | 4405 | hCoV-19/England/QEUH-B823C6/2020\|EPI_ISL_668242\|20 | ···················· | ···················· | ····················· | ···················· | ···················· | ···················· |
| T | 4406 | hCoV-19/England/QEUH-B7F371/2020\|EPI_ISL_668243\|20 | ···················· | ···················· | ····················· | ···················· | ···················· | ···················· |
| T | 4407 | hCoV-19/England/QEUH-B7FD82/2020\|EPI_ISL_668245\|20 | ···················· | ···················· | ····················· | ···················· | ···················· | ···················· |
| T | 4408 | hCoV-19/England/QEUH-B81488/2020\|EPI_ISL_668248\|20 | ···················· | ···················· | ····················· | ···················· | ···················· | ···················· |
| T | 4409 | hCoV-19/England/QEUH-B7C392/2020\|EPI_ISL_668249\|20 | ···················· | ···················· | ····················· | ···················· | ···················· | ···················· |
| T | 4410 | hCoV-19/England/CAMC-B77D3F/2020\|EPI_ISL_668251\|20 | ···················· | ···················· | ····················· | ···················· | ···················· | ···················· |
| T | 4411 | hCoV-19/England/QEUH-B7E8DD/2020\|EPI_ISL_668253\|20 | ···················· | ···················· | ····················· | ···················· | ···················· | ···················· |
| T | 4412 | hCoV-19/England/CAMC-B77D4E/2020\|EPI_ISL_668254\|20 | ···················· | ···················· | ····················· | ···················· | ···················· | ···················· |
| T | 4413 | hCoV-19/England/QEUH-B80692/2020\|EPI_ISL_668256\|20 | ···················· | ···················· | ····················· | ···················· | ···················· | ···················· |
| T | 4414 | hCoV-19/England/QEUH-B7C0C8/2020\|EPI_ISL_668257\|20 | ···················· | ···················· | ····················· | ···················· | ···················· | ···················· |
| T | 4415 | hCoV-19/England/QEUH-B7F61E/2020\|EPI_ISL_668258\|20 | ···················· | ···················· | ····················· | ···················· | ···················· | ···················· |
| T | 4416 | hCoV-19/England/QEUH-B7C15C/2020\|EPI_ISL_668259\|20 | ···················· | ···················· | ····················· | ···················· | ···················· | ···················· |
| T | 4417 | hCoV-19/England/QEUH-B82144/2020\|EPI_ISL_668260\|20 | ···················· | ···················· | ····················· | ···················· | ···················· | ···················· |
| T | 4418 | hCoV-19/England/QEUH-B7E3BE/2020\|EPI_ISL_668264\|20 | ···················· | ···················· | ····················· | ···················· | ···················· | ···················· |
| T | 4419 | hCoV-19/England/CAMC-B7783E/2020\|EPI_ISL_668265\|20 | ···················· | ···················· | ····················· | ···················· | ···················· | ···················· |
| T | 4420 | hCoV-19/England/QEUH-B80683/2020\|EPI_ISL_668266\|20 | ···················· | ···················· | ····················· | ···················· | ···················· | ···················· |
| T | 4421 | hCoV-19/England/QEUH-B814A6/2020\|EPI_ISL_668269\|20 | ···················· | ···················· | ····················· | ···················· | ···················· | ···················· |
| T | 4422 | hCoV-19/England/QEUH-B7E442/2020\|EPI_ISL_668271\|20 | ···················· | ···················· | ····················· | ···················· | ···················· | ···················· |
| T | 4423 | hCoV-19/England/QEUH-B7F6E1/2020\|EPI_ISL_668272\|20 | ···················· | ···················· | ····················· | ···················· | ···················· | ···················· |
| T | 4424 | hCoV-19/England/QEUH-B7F99D/2020\|EPI_ISL_668273\|20 | ···················· | ···················· | ····················· | ···················· | ···················· | ···················· |
| T | 4425 | hCoV-19/England/QEUH-B7C0E6/2020\|EPI_ISL_668276\|20 | ···················· | ·A·················· | ····················· | ···················· | ···················· | ···················· |
| T | 4426 | hCoV-19/England/QEUH-B7FD19/2020\|EPI_ISL_668277\|20 | ···················· | ·A·················· | ····················· | ···················· | ···················· | ···················· |
| T | 4427 | hCoV-19/England/QEUH-B813F4/2020\|EPI_ISL_668278\|20 | ···················· | ···················· | ····················· | ···················· | ······T············· | ···················· |
| T | 4428 | hCoV-19/England/QEUH-B7F9BB/2020\|EPI_ISL_668282\|20 | ···················· | ···················· | ····················· | ···················· | ···················· | ···················· |
| T | 4429 | hCoV-19/England/QEUH-B8241E/2020\|EPI_ISL_668284\|20 | ···················· | ···················· | ····················· | ···················· | ···················· | ···················· |
| T | 4430 | hCoV-19/England/QEUH-B7E47F/2020\|EPI_ISL_668286\|20 | ···················· | ·A·················· | ····················· | ···················· | ···················· | ···················· |
| T | 4431 | hCoV-19/England/QEUH-B7C110/2020\|EPI_ISL_668287\|20 | ···················· | ···················· | ····················· | ···················· | ···················· | ···················· |
| T | 4432 | hCoV-19/England/QEUH-B8145B/2020\|EPI_ISL_668288\|20 | ···················· | ···················· | ····················· | ···················· | ···················· | ···················· |
| T | 4433 | hCoV-19/England/QEUH-B8142E/2020\|EPI_ISL_668290\|20 | ···················· | ···················· | ····················· | ···················· | ···················· | ···················· |
| T | 4434 | hCoV-19/England/QEUH-B7F6B4/2020\|EPI_ISL_668291\|20 | ···················· | ···················· | ····················· | ···················· | ···················· | ···················· |
| T | 4435 | hCoV-19/England/CAMC-B7769E/2020\|EPI_ISL_668296\|20 | ···················· | ···················· | ····················· | ···················· | ···················· | ···················· |
| T | 4436 | hCoV-19/England/CAMC-B7782F/2020\|EPI_ISL_668299\|20 | ···················· | ···················· | ····················· | ···················· | ···················· | ···················· |
| T | 4437 | hCoV-19/England/QEUH-B7E8CE/2020\|EPI_ISL_668300\|20 | ···················· | ···················· | ····················· | ···················· | ···················· | ···················· |
| T | 4438 | hCoV-19/England/QEUH-B805FF/2020\|EPI_ISL_668303\|20 | ···················· | ···················· | ····················· | ···················· | ···················· | ···················· |
| T | 4439 | hCoV-19/England/QEUH-B7C365/2020\|EPI_ISL_668307\|20 | ···················· | ···················· | ····················· | ···················· | ···················· | ···················· |
| T | 4440 | hCoV-19/England/QEUH-B7BF80/2020\|EPI_ISL_668311\|20 | ···················· | ···················· | ····················· | ···················· | ···················· | ···················· |
| T | 4441 | hCoV-19/England/CAMC-B77625/2020\|EPI_ISL_668316\|20 | ···················· | ·A·················· | ····················· | ···················· | ···················· | ··A················· |
| T | 4442 | hCoV-19/England/CAMC-B7776E/2020\|EPI_ISL_668319\|20 | ···················· | ···················· | ····················· | ···················· | ···················· | ···················· |
| T | 4443 | hCoV-19/England/QEUH-B7E3DC/2020\|EPI_ISL_668320\|20 | ···················· | ···················· | ····················· | ···················· | ···················· | ···················· |
| T | 4444 | hCoV-19/England/QEUH-B7C3B0/2020\|EPI_ISL_668321\|20 | ···················· | ···················· | ····················· | ···················· | ···················· | ···················· |
| T | 4446 | hCoV-19/England/QEUH-B7F906/2020\|EPI_ISL_668328\|20 | ···················· | ···················· | ····················· | ···················· | ···················· | ···················· |
| T | 4447 | hCoV-19/England/CAMC-B77704/2020\|EPI_ISL_668336\|20 | ···················· | ···················· | ····················· | ···················· | ···················· | ···················· |
| T | 4448 | hCoV-19/England/QEUH-B820FC/2020\|EPI_ISL_668338\|20 | ···················· | ···················· | ····················· | ···················· | ···················· | ···················· |
| T | 4449 | hCoV-19/England/QEUH-B7E819/2020\|EPI_ISL_668339\|20 | ···················· | ···················· | ····················· | ···················· | ···················· | ···················· |
| T | 4450 | hCoV-19/Denmark/DCGC-10556/2020\|EPI_ISL_669596\|202 | ···················· | ···················· | ····················· | ···················· | ···················· | ···················· |
| T | 4451 | hCoV-19/Denmark/DCGC-10560/2020\|EPI_ISL_669597\|202 | ···················· | ···················· | ····················· | ···················· | ···················· | ···················· |
| T | 4452 | hCoV-19/Denmark/DCGC-10889/2020\|EPI_ISL_669598\|202 | ···················· | ···················· | ····················· | ···················· | ···················· | ···················· |
| T | 4453 | hCoV-19/Denmark/DCGC-11068/2020\|EPI_ISL_669599\|202 | ···················· | ···················· | ····················· | ···················· | ···················· | ···················· |

| | | | | | | | |
|---|---|---|---|---|---|---|---|
| T | 4454 hCoV-19/Denmark/DCGC-10215/2020\|EPI_ISL_669636\|202 | ···················· | ···················· | ····················· | ···················· | ···················· | ···················· |
| T | 4455 hCoV-19/Denmark/DCGC-10225/2020\|EPI_ISL_669637\|202 | ···················· | ···················· | ····················· | ···················· | ···················· | ···················· |
| T | 4456 hCoV-19/Denmark/DCGC-10240/2020\|EPI_ISL_669638\|202 | ···················· | ···················· | ····················· | ···················· | ···················· | ···················· |
| T | 4457 hCoV-19/Denmark/DCGC-10251/2020\|EPI_ISL_669639\|202 | ···················· | ···················· | ····················· | ···················· | ···················· | ···················· |
| T | 4458 hCoV-19/Denmark/DCGC-10269/2020\|EPI_ISL_669640\|202 | ···················· | ···················· | ····················· | ···················· | ···················· | ···················· |
| T | 4459 hCoV-19/Denmark/DCGC-10286/2020\|EPI_ISL_669641\|202 | ···················· | ···················· | ····················· | ···················· | ···················· | ···················· |
| T | 4460 hCoV-19/Denmark/DCGC-10288/2020\|EPI_ISL_669642\|202 | ···················· | ···················· | ····················· | ···················· | ···················· | ···················· |
| T | 4461 hCoV-19/Denmark/DCGC-10291/2020\|EPI_ISL_669643\|202 | ···················· | ···················· | ····················· | ···················· | ···················· | ···················· |
| T | 4462 hCoV-19/Denmark/DCGC-10307/2020\|EPI_ISL_669644\|202 | ···················· | ···················· | ····················· | ···················· | ···················· | ···················· |
| T | 4463 hCoV-19/Denmark/DCGC-10315/2020\|EPI_ISL_669645\|202 | ···················· | ···················· | ····················· | ···················· | ···················· | ···················· |
| T | 4464 hCoV-19/Denmark/DCGC-10321/2020\|EPI_ISL_669646\|202 | ···················· | ···················· | ····················· | ···················· | ···················· | ···················· |
| T | 4465 hCoV-19/Denmark/DCGC-10333/2020\|EPI_ISL_669647\|202 | ···················· | ···················· | ····················· | ···················· | ···················· | ···················· |
| T | 4466 hCoV-19/Denmark/DCGC-10335/2020\|EPI_ISL_669648\|202 | ···················· | ···················· | ····················· | ···················· | ···················· | ···················· |
| T | 4467 hCoV-19/Denmark/DCGC-10337/2020\|EPI_ISL_669649\|202 | ···················· | ···················· | ····················· | ···················· | ···················· | ···················· |
| T | 4468 hCoV-19/Denmark/DCGC-10349/2020\|EPI_ISL_669650\|202 | ···················· | ···················· | ····················· | ···················· | ···················· | ···················· |
| T | 4469 hCoV-19/Denmark/DCGC-10355/2020\|EPI_ISL_669651\|202 | ···················· | ···················· | ····················· | ···················· | ···················· | ···················· |
| T | 4470 hCoV-19/Denmark/DCGC-10365/2020\|EPI_ISL_669652\|202 | ···················· | ···················· | ····················· | ···················· | ···················· | ···················· |
| T | 4471 hCoV-19/Denmark/DCGC-10368/2020\|EPI_ISL_669653\|202 | ···················· | ···················· | ····················· | ···················· | ···················· | ···················· |
| T | 4472 hCoV-19/Denmark/DCGC-10371/2020\|EPI_ISL_669654\|202 | ···················· | ···················· | ····················· | ···················· | ···················· | ···················· |
| T | 4473 hCoV-19/Denmark/DCGC-10372/2020\|EPI_ISL_669655\|202 | ···················· | ···················· | ····················· | ···················· | ···················· | ···················· |
| T | 4474 hCoV-19/Denmark/DCGC-10377/2020\|EPI_ISL_669656\|202 | ···················· | ···················· | ····················· | ···················· | ···················· | ···················· |
| T | 4475 hCoV-19/Denmark/DCGC-10383/2020\|EPI_ISL_669657\|202 | ···················· | ···················· | ····················· | ···················· | ···················· | ···················· |
| T | 4476 hCoV-19/Denmark/DCGC-10392/2020\|EPI_ISL_669658\|202 | ···················· | ···················· | ····················· | ···················· | ···················· | ···················· |
| T | 4477 hCoV-19/Denmark/DCGC-10404/2020\|EPI_ISL_669659\|202 | ···················· | ···················· | ····················· | ···················· | ···················· | ···················· |
| T | 4478 hCoV-19/Denmark/DCGC-10469/2020\|EPI_ISL_669667\|202 | ···················· | ···················· | ····················· | ···················· | ···················· | ···················· |
| T | 4479 hCoV-19/Denmark/DCGC-10472/2020\|EPI_ISL_669668\|202 | ···················· | ···················· | ····················· | ···················· | ···················· | ···················· |
| T | 4480 hCoV-19/Denmark/DCGC-10485/2020\|EPI_ISL_669669\|202 | ···················· | ···················· | ····················· | ···················· | ···················· | ···················· |
| T | 4481 hCoV-19/Denmark/DCGC-11226/2020\|EPI_ISL_670552\|202 | ···················· | ···················· | ····················· | ···················· | ···················· | ···················· |
| T | 4482 hCoV-19/Denmark/DCGC-11525/2020\|EPI_ISL_670553\|202 | ···················· | ···················· | ····················· | ···················· | ···················· | ···················· |
| T | 4483 hCoV-19/Denmark/DCGC-11579/2020\|EPI_ISL_670554\|202 | ···················· | ···················· | ····················· | ···················· | ···················· | ···················· |
| T | 4484 hCoV-19/Denmark/DCGC-11600/2020\|EPI_ISL_670555\|202 | ···················· | ···················· | ····················· | ···················· | ···················· | ···················· |
| T | 4485 hCoV-19/Denmark/DCGC-11678/2020\|EPI_ISL_670556\|202 | ···················· | ···················· | ····················· | ···················· | ···················· | ···················· |
| T | 4486 hCoV-19/Denmark/DCGC-11716/2020\|EPI_ISL_670557\|202 | ···················· | ···················· | ····················· | ···················· | ···················· | ···················· |
| T | 4487 hCoV-19/Denmark/DCGC-11213/2020\|EPI_ISL_670576\|202 | ···················· | ···················· | ····················· | ···················· | ···················· | ···················· |
| T | 4488 hCoV-19/Denmark/DCGC-11223/2020\|EPI_ISL_670577\|202 | ···················· | ···················· | ····················· | ···················· | ···················· | ···················· |
| T | 4489 hCoV-19/Denmark/DCGC-11225/2020\|EPI_ISL_670578\|202 | ···················· | ···················· | ····················· | ···················· | ···················· | ···················· |
| T | 4490 hCoV-19/Denmark/DCGC-11239/2020\|EPI_ISL_670579\|202 | ···················· | ···················· | ····················· | ···················· | ···················· | ···················· |
| T | 4491 hCoV-19/Denmark/DCGC-11242/2020\|EPI_ISL_670580\|202 | ···················· | ·A·················· | ····················· | ···················· | ···················· | ···················· |
| T | 4492 hCoV-19/Denmark/DCGC-11245/2020\|EPI_ISL_670581\|202 | ···················· | ···················· | ····················· | ···················· | ···················· | ···················· |
| T | 4493 hCoV-19/Denmark/DCGC-11250/2020\|EPI_ISL_670582\|202 | ···················· | ···················· | ····················· | ···················· | ···················· | ···················· |
| T | 4494 hCoV-19/Denmark/DCGC-11253/2020\|EPI_ISL_670583\|202 | ···················· | ···················· | ····················· | ···················· | ···················· | ···················· |
| T | 4495 hCoV-19/Denmark/DCGC-11263/2020\|EPI_ISL_670584\|202 | ···················· | ···················· | ····················· | ···················· | ···················· | ···················· |
| T | 4496 hCoV-19/Denmark/DCGC-11264/2020\|EPI_ISL_670585\|202 | ···················· | ···················· | ····················· | ···················· | ···················· | ···················· |
| T | 4497 hCoV-19/Denmark/DCGC-11266/2020\|EPI_ISL_670586\|202 | ···················· | ···················· | ····················· | ···················· | ···················· | ···················· |
| T | 4498 hCoV-19/Denmark/DCGC-11271/2020\|EPI_ISL_670587\|202 | ···················· | ···················· | ····················· | ···················· | ···················· | ···················· |
| T | 4499 hCoV-19/Denmark/DCGC-11279/2020\|EPI_ISL_670588\|202 | ···················· | ···················· | ····················· | ···················· | ···················· | ···················· |
| T | 4500 hCoV-19/Denmark/DCGC-11284/2020\|EPI_ISL_670589\|202 | ···················· | ···················· | ····················· | ···················· | ···················· | ···················· |
| T | 4501 hCoV-19/Denmark/DCGC-11285/2020\|EPI_ISL_670590\|202 | ···················· | ···················· | ····················· | ···················· | ···················· | ···················· |
| T | 4502 hCoV-19/Denmark/DCGC-11311/2020\|EPI_ISL_670591\|202 | ···················· | ···················· | ····················· | ···················· | ···················· | ···················· |
| T | 4503 hCoV-19/Denmark/DCGC-11313/2020\|EPI_ISL_670592\|202 | ···················· | ···················· | ····················· | ···················· | ···················· | ···················· |
| T | 4504 hCoV-19/Denmark/DCGC-11315/2020\|EPI_ISL_670593\|202 | ···················· | ···················· | ····················· | ···················· | ···················· | ···················· |
| T | 4505 hCoV-19/Denmark/DCGC-11328/2020\|EPI_ISL_670594\|202 | ···················· | ···················· | ····················· | ···················· | ···················· | ···················· |
| T | 4506 hCoV-19/Denmark/DCGC-11329/2020\|EPI_ISL_670595\|202 | ···················· | ·A·················· | ····················· | ···················· | ···················· | ···················· |
| T | 4507 hCoV-19/Denmark/DCGC-11333/2020\|EPI_ISL_670596\|202 | ···················· | ···················· | ····················· | ···················· | ···················· | ···················· |
| T | 4508 hCoV-19/Denmark/DCGC-11346/2020\|EPI_ISL_670597\|202 | ···················· | ···················· | ····················· | ···················· | ···················· | ···················· |
| T | 4509 hCoV-19/Denmark/DCGC-11360/2020\|EPI_ISL_670598\|202 | ···················· | ···················· | ····················· | ···················· | ···················· | ···················· |
| T | 4510 hCoV-19/Denmark/DCGC-11368/2020\|EPI_ISL_670599\|202 | ···················· | ···················· | ····················· | ···················· | ···················· | ···················· |
| T | 4511 hCoV-19/Denmark/DCGC-11375/2020\|EPI_ISL_670600\|202 | ···················· | ···················· | ····················· | ···················· | ···················· | ···················· |
| T | 4512 hCoV-19/Denmark/DCGC-11391/2020\|EPI_ISL_670601\|202 | ···················· | ···················· | ····················· | ···················· | ···················· | ···················· |
| T | 4513 hCoV-19/Denmark/DCGC-11393/2020\|EPI_ISL_670602\|202 | ···················· | ···················· | ····················· | ···················· | ···················· | ···················· |
| T | 4514 hCoV-19/Denmark/DCGC-11421/2020\|EPI_ISL_670603\|202 | ···················· | ···················· | ····················· | ···················· | ···················· | ···················· |
| T | 4515 hCoV-19/Denmark/DCGC-11426/2020\|EPI_ISL_670604\|202 | ···················· | ···················· | ····················· | ···················· | ···················· | ···················· |
| T | 4516 hCoV-19/Denmark/DCGC-11438/2020\|EPI_ISL_670605\|202 | ···················· | ···················· | ····················· | ···················· | ···················· | ···················· |
| T | 4517 hCoV-19/Denmark/DCGC-11448/2020\|EPI_ISL_670606\|202 | ···················· | ···················· | ····················· | ···················· | ···················· | ···················· |
| T | 4518 hCoV-19/Denmark/DCGC-11454/2020\|EPI_ISL_670607\|202 | ···················· | ···················· | ····················· | ···················· | ···················· | ···················· |
| T | 4519 hCoV-19/Denmark/DCGC-11457/2020\|EPI_ISL_670608\|202 | ···················· | ···················· | ····················· | ···················· | ···················· | ···················· |
| T | 4520 hCoV-19/Denmark/DCGC-11458/2020\|EPI_ISL_670609\|202 | ···················· | ···················· | ····················· | ···················· | ···················· | ···················· |
| T | 4521 hCoV-19/Denmark/DCGC-11459/2020\|EPI_ISL_670610\|202 | ···················· | ···················· | ····················· | ···················· | ···················· | ···················· |
| T | 4522 hCoV-19/Denmark/DCGC-11460/2020\|EPI_ISL_670611\|202 | ···················· | ···················· | ····················· | ···················· | ···················· | ···················· |
| T | 4523 hCoV-19/Denmark/DCGC-11462/2020\|EPI_ISL_670612\|202 | ···················· | ···················· | ····················· | ···················· | ···················· | ···················· |
| T | 4524 hCoV-19/Denmark/DCGC-11473/2020\|EPI_ISL_670613\|202 | ···················· | ···················· | ····················· | ···················· | ···················· | ···················· |
| T | 4525 hCoV-19/Denmark/DCGC-11507/2020\|EPI_ISL_670614\|202 | ···················· | ···················· | ····················· | ···················· | ···················· | ···················· |
| T | 4526 hCoV-19/Denmark/DCGC-11520/2020\|EPI_ISL_670615\|202 | ···················· | ···················· | ····················· | ···················· | ···················· | ···················· |
| T | 4527 hCoV-19/Denmark/DCGC-11534/2020\|EPI_ISL_670616\|202 | ···················· | ···················· | ····················· | ···················· | ···················· | ···················· |
| T | 4528 hCoV-19/Denmark/DCGC-11539/2020\|EPI_ISL_670617\|202 | ···················· | ···················· | ····················· | ···················· | ···················· | ···················· |
| T | 4529 hCoV-19/Denmark/DCGC-11546/2020\|EPI_ISL_670618\|202 | ···················· | ···················· | ····················· | ···················· | ···················· | ···················· |
| T | 4530 hCoV-19/Denmark/DCGC-11557/2020\|EPI_ISL_670619\|202 | ···················· | ···················· | ····················· | ···················· | ···················· | ···················· |
| T | 4531 hCoV-19/Denmark/DCGC-11572/2020\|EPI_ISL_670620\|202 | ···················· | ···················· | ····················· | ···················· | ···················· | ···················· |
| T | 4532 hCoV-19/Denmark/DCGC-11574/2020\|EPI_ISL_670621\|202 | ···················· | ···················· | ····················· | ···················· | ···················· | ···················· |
| T | 4533 hCoV-19/Denmark/DCGC-11581/2020\|EPI_ISL_670622\|202 | ···················· | ···················· | ····················· | ···················· | ···················· | ···················· |

| | | | | | | | | |
|---|---|---|---|---|---|---|---|---|
| T | 4534 | hCoV-19/Denmark/DCGC-11588/2020\|EPI_ISL_670623\|202 | .................... | .................... | ..................... | .................... | .................... | .................... |
| T | 4535 | hCoV-19/Denmark/DCGC-11599/2020\|EPI_ISL_670624\|202 | .................... | .................... | ..................... | .................... | .................... | .................... |
| T | 4536 | hCoV-19/Denmark/DCGC-11628/2020\|EPI_ISL_670625\|202 | .................... | .................... | ..................... | .................... | .................... | .................... |
| T | 4537 | hCoV-19/Denmark/DCGC-11635/2020\|EPI_ISL_670626\|202 | .................... | .................... | ..................... | .................... | .................... | .................... |
| T | 4538 | hCoV-19/Denmark/DCGC-11637/2020\|EPI_ISL_670627\|202 | .................... | .................... | ..................... | .................... | .................... | .................... |
| T | 4539 | hCoV-19/Denmark/DCGC-11638/2020\|EPI_ISL_670628\|202 | .................... | .................... | ..................... | .................... | .................... | .................... |
| T | 4540 | hCoV-19/Denmark/DCGC-11646/2020\|EPI_ISL_670629\|202 | .................... | .................... | ..................... | .................... | .................... | .................... |
| T | 4541 | hCoV-19/Denmark/DCGC-11654/2020\|EPI_ISL_670630\|202 | .................... | .................... | ..................... | .................... | .................... | .................... |
| T | 4542 | hCoV-19/Denmark/DCGC-11663/2020\|EPI_ISL_670631\|202 | .................... | .................... | ..................... | .................... | .................... | .................... |
| T | 4543 | hCoV-19/Denmark/DCGC-11676/2020\|EPI_ISL_670632\|202 | .................... | .................... | ..................... | .................... | .................... | .................... |
| T | 4544 | hCoV-19/Denmark/DCGC-11696/2020\|EPI_ISL_670633\|202 | .................... | .................... | ..................... | .................... | .................... | .................... |
| T | 4545 | hCoV-19/Denmark/DCGC-11718/2020\|EPI_ISL_670634\|202 | .................... | .A.................. | ..................... | .................... | .................... | .................... |
| T | 4546 | hCoV-19/Denmark/DCGC-11768/2020\|EPI_ISL_670635\|202 | .................... | .................... | ..................... | .................... | .................... | .................... |
| T | 4547 | hCoV-19/Denmark/DCGC-11781/2020\|EPI_ISL_670636\|202 | .................... | .................... | ..................... | .................... | .................... | .................... |
| T | 4548 | hCoV-19/Denmark/DCGC-11786/2020\|EPI_ISL_670637\|202 | .................... | .................... | ..................... | .................... | .................... | .................... |
| T | 4549 | hCoV-19/Denmark/DCGC-11810/2020\|EPI_ISL_670638\|202 | .................... | .................... | ..................... | .................... | .................... | .................... |
| T | 4550 | hCoV-19/Denmark/DCGC-11833/2020\|EPI_ISL_670639\|202 | .................... | .................... | ..................... | .................... | .................... | .................... |
| T | 4551 | hCoV-19/Ireland/CO-NVRL-75IRL84568/2020\|EPI_ISL_67 | .................... | .................... | ..................... | .................... | .................... | .................... |
| T | 4552 | hCoV-19/Ireland/D-NVRL-20IRL87996/2020\|EPI_ISL_671 | .................... | .................... | ..................... | .................... | .................... | .................... |
| T | 4553 | hCoV-19/Ireland/D-NVRL-75IRL26026/2020\|EPI_ISL_671 | .................... | .................... | ..................... | .................... | .................... | .................... |
| T | 4554 | hCoV-19/Ireland/D-NVRL-75IRL26146/2020\|EPI_ISL_671 | .................... | .A.................. | ..................... | .................... | .................... | ...A................ |
| T | 4555 | hCoV-19/Ireland/D-NVRL-75IRL67616/2020\|EPI_ISL_671 | .................... | .................... | ..................... | .................... | .................... | .................... |
| T | 4556 | hCoV-19/Ireland/D-NVRL-75IRL68622/2020\|EPI_ISL_671 | .................... | .................... | ..................... | .................... | .................... | .................... |
| T | 4557 | hCoV-19/Ireland/D-NVRL-75IRL75521/2020\|EPI_ISL_671 | .................... | .................... | ..................... | .................... | .................... | .................... |
| T | 4558 | hCoV-19/Ireland/D-NVRL-75IRL76096/2020\|EPI_ISL_671 | .................... | .................... | ..................... | .................... | .................... | .................... |
| T | 4559 | hCoV-19/Ireland/D-NVRL-75IRL77046/2020\|EPI_ISL_671 | .................... | .................... | ..................... | .................... | .................... | .................... |
| T | 4561 | hCoV-19/Ireland/D-NVRL-75IRL77394/2020\|EPI_ISL_671 | .................... | .................... | ..................... | .................... | .................... | .................... |
| T | 4562 | hCoV-19/Ireland/D-NVRL-75IRL88687/2020\|EPI_ISL_671 | .................... | .................... | ..................... | .................... | .................... | .................... |
| T | 4564 | hCoV-19/Ireland/LS-NVRL-75IRL70478/2020\|EPI_ISL_67 | .................... | .................... | ..................... | .................... | .................... | .................... |
| T | 4565 | hCoV-19/Ireland/LS-NVRL-75IRL79312/2020\|EPI_ISL_67 | .................... | .................... | ..................... | .................... | .................... | .................... |
| T | 4566 | hCoV-19/Ireland/LS-NVRL-75IRL79332/2020\|EPI_ISL_67 | .................... | .................... | ..................... | .................... | .................... | .................... |
| T | 4567 | hCoV-19/Ireland/MO-NVRL-75IRL73163/2020\|EPI_ISL_67 | .................... | .................... | ..................... | .................... | .................... | .................... |
| T | 4568 | hCoV-19/Ireland/OY-NVRL-75IRL34224/2020\|EPI_ISL_67 | .................... | .................... | ..................... | .................... | .................... | .................... |
| T | 4569 | hCoV-19/Ireland/OY-NVRL-75IRL70179/2020\|EPI_ISL_67 | .................... | .................... | ..................... | .................... | .................... | .................... |
| T | 4570 | hCoV-19/Ireland/OY-NVRL-75IRL90286/2020\|EPI_ISL_67 | .................... | .................... | ..................... | .................... | .................... | .................... |
| T | 4571 | hCoV-19/Ireland/RN-NVRL-75IRL23066/2020\|EPI_ISL_67 | .................... | .................... | ..................... | .................... | .................... | .................... |
| T | 4572 | hCoV-19/Ireland/RN-NVRL-75IRL84062/2020\|EPI_ISL_67 | .................... | .................... | ..................... | .................... | .................... | .................... |
| T | 4573 | hCoV-19/Ireland/SO-NVRL-75IRL23335/2020\|EPI_ISL_67 | .................... | .................... | ..................... | .................... | .................... | .................... |
| T | 4574 | hCoV-19/Ireland/TA-NVRL-75IRL33036/2020\|EPI_ISL_67 | .................... | .................... | ..................... | .................... | .................... | .................... |
| T | 4575 | hCoV-19/Ireland/WH-NVRL-75IRL28485/2020\|EPI_ISL_67 | .................... | .................... | ..................... | .................... | .................... | .................... |
| T | 4576 | hCoV-19/Ireland/WH-NVRL-75IRL69800/2020\|EPI_ISL_67 | .................... | .................... | ..................... | .................... | .................... | .................... |
| T | 4577 | hCoV-19/Ireland/WH-NVRL-75IRL78270/2020\|EPI_ISL_67 | .................... | .................... | ..................... | .................... | .................... | .................... |
| T | 4578 | hCoV-19/Ireland/WX-NVRL-75IRL33947/2020\|EPI_ISL_67 | .................... | .................... | ..................... | .................... | .................... | .................... |
| T | 4579 | hCoV-19/Ireland/WX-NVRL-75IRL33999/2020\|EPI_ISL_67 | .................... | .................... | ..................... | .................... | .................... | .................... |
| T | 4580 | hCoV-19/France/OCC-94/2020\|EPI_ISL_671941\|2020-09- | .................... | .................... | ..................... | .................... | .................... | .................... |
| T | 4581 | hCoV-19/France/OCC-95/2020\|EPI_ISL_671942\|2020-09- | .................... | .................... | ..................... | .................... | .................... | .................... |
| T | 4582 | hCoV-19/France/OCC-96/2020\|EPI_ISL_671943\|2020-07- | .................... | .................... | ..................... | .................... | .................... | .................... |
| T | 4583 | hCoV-19/France/OCC-97/2020\|EPI_ISL_671944\|2020-10- | .................... | .................... | ..................... | .................... | .................... | .................... |
| T | 4584 | hCoV-19/France/OCC-98/2020\|EPI_ISL_671945\|2020-10- | .................... | .................... | ..................... | .................... | .................... | .................... |
| T | 4585 | hCoV-19/France/OCC-100/2020\|EPI_ISL_671946\|2020-10 | .................... | .................... | ..................... | .................... | .................... | .................... |
| T | 4586 | hCoV-19/France/OCC-101/2020\|EPI_ISL_671947\|2020-10 | .................... | .................... | ..................... | .................... | .................... | .................... |
| T | 4587 | hCoV-19/France/OCC-102/2020\|EPI_ISL_671948\|2020-10 | .................... | .................... | ..................... | .................... | .................... | .................... |
| T | 4588 | hCoV-19/France/OCC-103/2020\|EPI_ISL_671949\|2020-10 | .................... | .................... | ..................... | .................... | .................... | .................... |
| T | 4589 | hCoV-19/France/OCC-104/2020\|EPI_ISL_671950\|2020-10 | .................... | .................... | ..................... | .................... | .................... | .................... |
| T | 4590 | hCoV-19/France/OCC-105/2020\|EPI_ISL_671951\|2020-10 | .................... | .................... | ..................... | .................... | .................... | .................... |
| T | 4591 | hCoV-19/France/OCC-106/2020\|EPI_ISL_671952\|2020-10 | .................... | .................... | ..................... | .................... | .................... | .................... |
| T | 4592 | hCoV-19/France/OCC-107/2020\|EPI_ISL_671953\|2020-10 | .................... | .................... | ..................... | .................... | .................... | .................... |
| T | 4593 | hCoV-19/France/OCC-108/2020\|EPI_ISL_671954\|2020-10 | .................... | .................... | ..................... | .................... | .................... | .................... |
| T | 4594 | hCoV-19/France/OCC-109/2020\|EPI_ISL_671955\|2020-10 | .................... | .................... | ..................... | .................... | .................... | .................... |
| T | 4595 | hCoV-19/France/OCC-110/2020\|EPI_ISL_671956\|2020-10 | .................... | .................... | ..................... | .................... | .................... | .................... |
| T | 4596 | hCoV-19/France/OCC-111/2020\|EPI_ISL_671957\|2020-10 | .................... | .................... | ..................... | .................... | .................... | .................... |
| T | 4597 | hCoV-19/France/OCC-112/2020\|EPI_ISL_671958\|2020-10 | .................... | .................... | ..................... | .................... | .................... | .................... |
| T | 4598 | hCoV-19/France/OCC-113/2020\|EPI_ISL_671959\|2020-10 | .................... | .................... | ..................... | .................... | .................... | .................... |
| T | 4599 | hCoV-19/France/OCC-115/2020\|EPI_ISL_671960\|2020-10 | .................... | .................... | ..................... | .................... | .................... | .................... |
| T | 4600 | hCoV-19/France/OCC-116/2020\|EPI_ISL_671961\|2020-10 | .................... | .................... | ..................... | .................... | .................... | .................... |
| T | 4601 | hCoV-19/France/OCC-117/2020\|EPI_ISL_671962\|2020-10 | .................... | .................... | ..................... | .................... | .................... | .................... |
| T | 4602 | hCoV-19/France/OCC-118/2020\|EPI_ISL_671963\|2020-10 | .................... | .................... | ..................... | .................... | .................... | .................... |
| T | 4603 | hCoV-19/France/OCC-119/2020\|EPI_ISL_671964\|2020-09 | .................... | .................... | ..................... | .................... | .................... | .................... |
| T | 4604 | hCoV-19/France/OCC-120/2020\|EPI_ISL_671965\|2020-10 | .................... | .................... | ..................... | .................... | .................... | .................... |
| T | 4605 | hCoV-19/France/OCC-121/2020\|EPI_ISL_671966\|2020-10 | .................... | .................... | ..................... | .................... | .................... | .................... |
| T | 4606 | hCoV-19/France/OCC-122/2020\|EPI_ISL_671967\|2020-10 | .................... | .................... | ..................... | .................... | .................... | .................... |
| T | 4607 | hCoV-19/France/OCC-123/2020\|EPI_ISL_671968\|2020-08 | .................... | .................... | ..................... | .................... | .................... | .................... |
| T | 4608 | hCoV-19/France/OCC-124/2020\|EPI_ISL_671969\|2020-10 | .................... | .................... | ..................... | .................... | .................... | .................... |
| T | 4609 | hCoV-19/England/QEUH-B6892E/2020\|EPI_ISL_662016\|20 | .................... | .................... | ..................... | .................... | .................... | .................... |
| T | 4610 | hCoV-19/England/ALDP-B74D41/2020\|EPI_ISL_662018\|20 | .................... | .................... | ..................... | .................... | .................... | .................... |
| T | 4611 | hCoV-19/England/ALDP-B75126/2020\|EPI_ISL_662019\|20 | .................... | .................... | ..................... | .................... | .................... | .................... |
| T | 4612 | hCoV-19/England/QEUH-B691E9/2020\|EPI_ISL_662021\|20 | .................... | .................... | ..................... | .................... | .................... | .................... |
| T | 4613 | hCoV-19/England/QEUH-B674C4/2020\|EPI_ISL_662024\|20 | .................... | .........A.......... | ..................... | .................... | ......T............. | .................... |
| T | 4614 | hCoV-19/England/ALDP-B755EE/2020\|EPI_ISL_662025\|20 | .................... | .................... | ..................... | .................... | .................... | .................... |
| T | 4615 | hCoV-19/England/QEUH-B68EA7/2020\|EPI_ISL_662027\|20 | .................... | .A.................. | ..................... | .................... | .................... | .................... |

| | | | | | | | | |
|---|---|---|---|---|---|---|---|---|
| T | 4616 | hCoV-19/England/QEUH-B686FA/2020\|EPI_ISL_662028\|20 | ···················· | ···················· | ···················· | ···················· | ···················· | ···················· |
| T | 4617 | hCoV-19/England/ALDP-B74330/2020\|EPI_ISL_662029\|20 | ···················· | ·A·················· | ···················· | ···················· | ···················· | ···················· |
| T | 4618 | hCoV-19/England/QEUH-B68663/2020\|EPI_ISL_662030\|20 | ···················· | ·A·················· | ···················· | ···················· | ···················· | ···················· |
| T | 4619 | hCoV-19/England/ALDP-B73FE2/2020\|EPI_ISL_662031\|20 | ···················· | ···················· | ···················· | ···················· | ···················· | ···················· |
| T | 4620 | hCoV-19/England/ALDP-B7527E/2020\|EPI_ISL_662032\|20 | ···················· | ···················· | ···················· | ···················· | ···················· | ···················· |
| T | 4621 | hCoV-19/England/QEUH-B67488/2020\|EPI_ISL_662035\|20 | ···················· | ········A··········· | ···················· | ···················· | ·····T·············· | ···················· |
| T | 4622 | hCoV-19/England/ALDP-B74312/2020\|EPI_ISL_662036\|20 | ···················· | ···················· | ···················· | ···················· | ···················· | ···················· |
| T | 4623 | hCoV-19/England/QEUH-B68EB6/2020\|EPI_ISL_662037\|20 | ···················· | ···················· | ···················· | ···················· | ···················· | ···················· |
| T | 4624 | hCoV-19/England/ALDP-B7528D/2020\|EPI_ISL_662038\|20 | ···················· | ···················· | ···················· | ···················· | ···················· | ···················· |
| T | 4625 | hCoV-19/England/QEUH-B69152/2020\|EPI_ISL_662039\|20 | ···················· | ···················· | ···················· | ···················· | ···················· | ···················· |
| T | 4626 | hCoV-19/England/QEUH-B6925F/2020\|EPI_ISL_662040\|20 | ···················· | ···················· | ···················· | ···················· | ···················· | ···················· |
| T | 4627 | hCoV-19/England/QEUH-B68DD7/2020\|EPI_ISL_662042\|20 | ···················· | ···················· | ···················· | ···················· | ···················· | ···················· |
| T | 4628 | hCoV-19/England/ALDP-B74D05/2020\|EPI_ISL_662043\|20 | ···················· | ···················· | ···················· | ···················· | ···················· | ···················· |
| T | 4629 | hCoV-19/England/QEUH-B69222/2020\|EPI_ISL_662044\|20 | ···················· | ···················· | ···················· | ···················· | ···················· | ···················· |
| T | 4630 | hCoV-19/England/ALDP-B747AD/2020\|EPI_ISL_662045\|20 | ···················· | ···················· | ···················· | ···················· | ···················· | ···················· |
| T | 4631 | hCoV-19/England/ALDP-B75250/2020\|EPI_ISL_662046\|20 | ···················· | ···················· | ···················· | ···················· | ···················· | ···················· |
| T | 4632 | hCoV-19/England/ALDP-B73FD3/2020\|EPI_ISL_662047\|20 | ···················· | ···················· | ···················· | ···················· | ···················· | ···················· |
| T | 4633 | hCoV-19/England/ALDP-B742D9/2020\|EPI_ISL_662048\|20 | ···················· | ···················· | ···················· | ···················· | ···················· | ···················· |
| T | 4634 | hCoV-19/England/ALDP-B74FD2/2020\|EPI_ISL_662049\|20 | ···················· | ···················· | ···················· | ···················· | ···················· | ···················· |
| T | 4635 | hCoV-19/England/ALDP-B74743/2020\|EPI_ISL_662051\|20 | ···················· | ···················· | ···················· | ···················· | ···················· | ···················· |
| T | 4636 | hCoV-19/England/QEUH-B68E2F/2020\|EPI_ISL_662052\|20 | ···················· | ···················· | ···················· | ···················· | ···················· | ···················· |
| T | 4637 | hCoV-19/England/ALDP-B75636/2020\|EPI_ISL_662053\|20 | ···················· | ···················· | ···················· | ···················· | ···················· | ···················· |
| T | 4638 | hCoV-19/England/QEUH-B6926E/2020\|EPI_ISL_662054\|20 | ···················· | ···················· | ···················· | ···················· | ···················· | ···················· |
| T | 4639 | hCoV-19/England/ALDP-B7497A/2020\|EPI_ISL_662055\|20 | ···················· | ···················· | ···················· | ···················· | ···················· | ···················· |
| T | 4640 | hCoV-19/England/ALDP-B7496B/2020\|EPI_ISL_662057\|20 | ···················· | ···················· | ···················· | ···················· | ···················· | ···················· |
| T | 4641 | hCoV-19/England/ALDP-B75654/2020\|EPI_ISL_662058\|20 | ···················· | ···················· | ···················· | ···················· | ···················· | ···················· |
| T | 4642 | hCoV-19/England/QEUH-B68DB9/2020\|EPI_ISL_662059\|20 | ···················· | ···················· | ···················· | ···················· | ···················· | ···················· |
| T | 4643 | hCoV-19/England/ALDP-B744F1/2020\|EPI_ISL_662060\|20 | ···················· | ···················· | ···················· | ···················· | ···················· | ···················· |
| T | 4644 | hCoV-19/England/ALDP-B7450D/2020\|EPI_ISL_662061\|20 | ···················· | ···················· | ···················· | ···················· | ···················· | ···················· |
| T | 4645 | hCoV-19/England/ALDP-B74D7E/2020\|EPI_ISL_662062\|20 | ···················· | ···················· | ···················· | ···················· | ···················· | ···················· |
| T | 4646 | hCoV-19/England/ALDP-B742BB/2020\|EPI_ISL_662063\|20 | ···················· | ···················· | ···················· | ···················· | ···················· | ···················· |
| T | 4647 | hCoV-19/England/QEUH-B6800B/2020\|EPI_ISL_662064\|20 | ···················· | ···················· | ···················· | ···················· | ···················· | ···················· |
| T | 4648 | hCoV-19/England/ALDP-B7538A/2020\|EPI_ISL_662065\|20 | ···················· | ···················· | ···················· | ···················· | ···················· | ···················· |
| T | 4649 | hCoV-19/England/QEUH-B66F1F/2020\|EPI_ISL_662066\|20 | ···················· | ···················· | ···················· | ···················· | ···················· | ···················· |
| T | 4650 | hCoV-19/England/ALDP-B74C80/2020\|EPI_ISL_662067\|20 | ···················· | ······A············· | ···················· | ···················· | ···················· | ···················· |
| T | 4651 | hCoV-19/England/ALDP-B7493E/2020\|EPI_ISL_662068\|20 | ···················· | ···················· | ···················· | ···················· | ···················· | ···················· |
| T | 4652 | hCoV-19/England/QEUH-B69213/2020\|EPI_ISL_662069\|20 | ···················· | ···················· | ···················· | ···················· | ···················· | ···················· |
| T | 4653 | hCoV-19/England/ALDP-B74DAB/2020\|EPI_ISL_662071\|20 | ···················· | ···················· | ···················· | ···················· | ···················· | ···················· |
| T | 4654 | hCoV-19/England/QEUH-B68627/2020\|EPI_ISL_662072\|20 | ···················· | ···················· | ···················· | ···················· | ···················· | ···················· |
| T | 4655 | hCoV-19/England/QEUH-B68742/2020\|EPI_ISL_662073\|20 | ···················· | ···················· | ···················· | ···················· | ···················· | ···················· |
| T | 4656 | hCoV-19/England/ALDP-B749D4/2020\|EPI_ISL_662074\|20 | ···················· | ···················· | ···················· | ···················· | ···················· | ···················· |
| T | 4657 | hCoV-19/England/QEUH-B68171/2020\|EPI_ISL_662075\|20 | ···················· | ···················· | ···················· | ···················· | ···················· | ···················· |
| T | 4658 | hCoV-19/England/QEUH-B69231/2020\|EPI_ISL_662076\|20 | ···················· | ···················· | ···················· | ···················· | ···················· | ···················· |
| T | 4659 | hCoV-19/England/QEUH-B66EA9/2020\|EPI_ISL_662077\|20 | ···················· | ···················· | ···················· | ···················· | ···················· | ···················· |
| T | 4660 | hCoV-19/England/ALDP-B7434F/2020\|EPI_ISL_662078\|20 | ···················· | ···················· | ···················· | ···················· | ···················· | ···················· |
| T | 4661 | hCoV-19/England/ALDP-B746FB/2020\|EPI_ISL_662079\|20 | ···················· | ···················· | ···················· | ···················· | ···················· | ···················· |
| T | 4662 | hCoV-19/England/ALDP-B7537B/2020\|EPI_ISL_662081\|20 | ···················· | ···················· | ···················· | ···················· | ···················· | ···················· |
| T | 4663 | hCoV-19/England/QEUH-B68074/2020\|EPI_ISL_662082\|20 | ···················· | ···················· | ···················· | ···················· | ···················· | ···················· |
| T | 4664 | hCoV-19/England/QEUH-B673A9/2020\|EPI_ISL_662083\|20 | ···················· | ···················· | ···················· | ···················· | ···················· | ···················· |
| T | 4665 | hCoV-19/England/QEUH-B686EB/2020\|EPI_ISL_662084\|20 | ···················· | ···················· | ···················· | ···················· | ···················· | ···················· |
| T | 4666 | hCoV-19/England/QEUH-B68180/2020\|EPI_ISL_662085\|20 | ···················· | ···················· | ···················· | ···················· | ···················· | ···················· |
| T | 4667 | hCoV-19/England/ALDP-B74725/2020\|EPI_ISL_662086\|20 | ···················· | ···················· | ···················· | ···················· | ···················· | ···················· |
| T | 4668 | hCoV-19/England/QEUH-B688E5/2020\|EPI_ISL_662087\|20 | ···················· | ···················· | ···················· | ···················· | ···················· | ···················· |
| T | 4669 | hCoV-19/England/QEUH-B66EB8/2020\|EPI_ISL_662088\|20 | ···················· | ···················· | ···················· | ···················· | ···················· | ···················· |
| T | 4670 | hCoV-19/England/QEUH-B68092/2020\|EPI_ISL_662089\|20 | ···················· | ···················· | ···················· | ···················· | ···················· | ···················· |
| T | 4671 | hCoV-19/England/QEUH-B68636/2020\|EPI_ISL_662090\|20 | ···················· | ···················· | ···················· | ···················· | ···················· | ···················· |
| T | 4672 | hCoV-19/England/QEUH-B68F95/2020\|EPI_ISL_662091\|20 | ···················· | ···················· | ···················· | ···················· | ···················· | ···················· |
| T | 4673 | hCoV-19/England/QEUH-B680DE/2020\|EPI_ISL_662092\|20 | ···················· | ···················· | ···················· | ···················· | ···················· | ···················· |
| T | 4674 | hCoV-19/England/ALDP-B752C9/2020\|EPI_ISL_662093\|20 | ···················· | ···················· | ···················· | ···················· | ···················· | ···················· |
| T | 4675 | hCoV-19/England/QEUH-B68F0E/2020\|EPI_ISL_662094\|20 | ···················· | ···················· | ···················· | ···················· | ···················· | ···················· |
| T | 4676 | hCoV-19/England/QEUH-B691AD/2020\|EPI_ISL_662096\|20 | ···················· | ···················· | ···················· | ···················· | ···················· | ···················· |
| T | 4677 | hCoV-19/England/ALDP-B74303/2020\|EPI_ISL_662098\|20 | ···················· | ·A·················· | ···················· | ···················· | ···················· | ··A················· |
| T | 4679 | hCoV-19/England/LOND-1250628/2020\|EPI_ISL_664742\|2 | ···················· | ···················· | ···················· | ···················· | ···················· | ···················· |
| T | 4680 | hCoV-19/England/ALDP-B73BEE/2020\|EPI_ISL_664747\|20 | ···················· | ···················· | ···················· | ···················· | ···················· | ···················· |
| T | 4681 | hCoV-19/England/CAMB-1BB7C2/2020\|EPI_ISL_664749\|20 | ···················· | ···················· | ···················· | ···················· | ···················· | ···················· |
| T | 4682 | hCoV-19/England/CAMB-1BB874/2020\|EPI_ISL_664750\|20 | ···················· | ···················· | ···················· | ···················· | ···················· | ···················· |
| T | 4683 | hCoV-19/England/CAMB-1BB892/2020\|EPI_ISL_664751\|20 | ···················· | ···················· | ···················· | ···················· | ···················· | ···················· |
| T | 4684 | hCoV-19/England/CAMC-B2335E/2020\|EPI_ISL_664752\|20 | ···················· | ···················· | ···················· | ···················· | ···················· | ···················· |
| T | 4685 | hCoV-19/England/LOND-12506CE/2020\|EPI_ISL_664753\|2 | ···················· | ···················· | ···················· | ···················· | ···················· | ···················· |
| T | 4687 | hCoV-19/England/NOTT-11C291/2020\|EPI_ISL_665901\|20 | ···················· | ···················· | ···················· | ···················· | ···················· | ···················· |
| T | 4688 | hCoV-19/England/NOTT-11C48C/2020\|EPI_ISL_665902\|20 | ···················· | ···················· | ···················· | ···················· | ···················· | ···················· |
| T | 4690 | hCoV-19/England/NOTT-11C923/2020\|EPI_ISL_665904\|20 | ···················· | ·A·················· | ···················· | ···················· | ···················· | ··A················· |
| T | 4691 | hCoV-19/England/NOTT-11C941/2020\|EPI_ISL_665905\|20 | ···················· | ···················· | ···················· | ···················· | ···················· | ···················· |
| T | 4692 | hCoV-19/England/NOTT-11CC66/2020\|EPI_ISL_665906\|20 | ···················· | ···················· | ···················· | ···················· | ···················· | ···················· |
| T | 4693 | hCoV-19/England/NOTT-11CD09/2020\|EPI_ISL_665907\|20 | ···················· | ···················· | ···················· | ···················· | ···················· | ···················· |
| T | 4694 | hCoV-19/England/NOTT-11CF03/2020\|EPI_ISL_665908\|20 | ···················· | ···················· | ···················· | ···················· | ···················· | ···················· |
| T | 4695 | hCoV-19/England/NOTT-11CF7C/2020\|EPI_ISL_665909\|20 | ···················· | ···················· | ···················· | ···················· | ···················· | ···················· |
| T | 4696 | hCoV-19/England/NOTT-11CFB8/2020\|EPI_ISL_665910\|20 | ···················· | ···················· | ···················· | ···················· | ···················· | ···················· |
| T | 4697 | hCoV-19/England/NOTT-11CFC7/2020\|EPI_ISL_665911\|20 | ···················· | ···················· | ···················· | ···················· | ···················· | ···················· |
| T | 4698 | hCoV-19/England/NOTT-11CFD6/2020\|EPI_ISL_665912\|20 | ···················· | ···················· | ···················· | ···················· | ···················· | ···················· |

| | | | | | | | | |
|---|---|---|---|---|---|---|---|---|
| T | 4699 | hCoV-19/England/NOTT-11CFF4/2020\|EPI_ISL_665913\|20 | ···················· | ···················· | ····················· | ···················· | ···················· | ···················· |
| T | 4700 | hCoV-19/England/NOTT-11D00F/2020\|EPI_ISL_665914\|20 | ···················· | ···················· | ····················· | ···················· | ···················· | ···················· |
| T | 4701 | hCoV-19/England/NOTT-11D03C/2020\|EPI_ISL_665915\|20 | ···················· | ···················· | ····················· | ···················· | ···················· | ···················· |
| T | 4702 | hCoV-19/England/NOTT-11D0B4/2020\|EPI_ISL_665916\|20 | ···················· | ···················· | ····················· | ···················· | ···················· | ···················· |
| T | 4703 | hCoV-19/England/NOTT-11D0C3/2020\|EPI_ISL_665917\|20 | ···················· | ···················· | ····················· | ···················· | ···················· | ···················· |
| T | 4704 | hCoV-19/England/NOTT-11D157/2020\|EPI_ISL_665918\|20 | ···················· | ···················· | ····················· | ···················· | ···················· | ···················· |
| T | 4705 | hCoV-19/England/NOTT-11D1EE/2020\|EPI_ISL_665919\|20 | ···················· | ···················· | ····················· | ···················· | ···················· | ···················· |
| T | 4706 | hCoV-19/England/QEUH-B7EBA7/2020\|EPI_ISL_668120\|20 | ···················· | ···················· | ····················· | ···················· | ···················· | ···················· |
| T | 4707 | hCoV-19/England/QEUH-B7EB2F/2020\|EPI_ISL_668121\|20 | ···················· | ···················· | ····················· | ···················· | ···················· | ···················· |
| T | 4708 | hCoV-19/England/CAMC-B79966/2020\|EPI_ISL_668122\|20 | ···················· | ···················· | ····················· | ···················· | ···················· | ···················· |
| T | 4709 | hCoV-19/England/QEUH-B7F2B0/2020\|EPI_ISL_668124\|20 | ···················· | ···················· | ····················· | ···················· | ···················· | ···················· |
| T | 4710 | hCoV-19/England/CAMC-B78E4A/2020\|EPI_ISL_668127\|20 | ···················· | ···················· | ····················· | ···················· | ···················· | ···················· |
| T | 4711 | hCoV-19/England/CAMC-B796D8/2020\|EPI_ISL_668128\|20 | ···················· | ···················· | ····················· | ···················· | ···················· | ···················· |
| T | 4712 | hCoV-19/England/QEUH-B7F9CA/2020\|EPI_ISL_668129\|20 | ···················· | ···················· | ····················· | ···················· | ···················· | ···················· |
| T | 4713 | hCoV-19/England/QEUH-B7C338/2020\|EPI_ISL_668130\|20 | ···················· | ···················· | ····················· | ···················· | ···················· | ···················· |
| T | 4714 | hCoV-19/England/QEUH-B7FD91/2020\|EPI_ISL_668132\|20 | ···················· | ···················· | ····················· | ···················· | ···················· | ···················· |
| T | 4715 | hCoV-19/England/QEUH-B8242D/2020\|EPI_ISL_668134\|20 | ···················· | ···················· | ····················· | ···················· | ···················· | ···················· |
| T | 4716 | hCoV-19/England/QEUH-B7E415/2020\|EPI_ISL_668135\|20 | ···················· | ···················· | ····················· | ···················· | ···················· | ···················· |
| T | 4717 | hCoV-19/England/CAMC-B7768F/2020\|EPI_ISL_668136\|20 | ···················· | ·A·················· | ····················· | ···················· | ···················· | ···A················ |
| T | 4718 | hCoV-19/England/CAMC-B776BC/2020\|EPI_ISL_668137\|20 | ···················· | ···················· | ····················· | ···················· | ···················· | ···················· |
| T | 4720 | hCoV-19/England/QEUH-B7F678/2020\|EPI_ISL_668144\|20 | ···················· | ···················· | ····················· | ···················· | ···················· | ···················· |
| T | 4721 | hCoV-19/England/QEUH-B8146A/2020\|EPI_ISL_668145\|20 | ···················· | ···················· | ····················· | ···················· | ···················· | ···················· |
| T | 4722 | hCoV-19/England/CAMC-B77FEE/2020\|EPI_ISL_668148\|20 | ···················· | ···················· | ····················· | ···················· | ···················· | ···················· |
| T | 4723 | hCoV-19/England/QEUH-B7BFCC/2020\|EPI_ISL_668150\|20 | ···················· | ···················· | ····················· | ···················· | ···················· | ···················· |
| T | 4724 | hCoV-19/England/QEUH-B7C1D4/2020\|EPI_ISL_668157\|20 | ···················· | ···················· | ····················· | ···················· | ······T············· | ···················· |
| T | 4725 | hCoV-19/England/QEUH-B7C0AA/2020\|EPI_ISL_668159\|20 | ···················· | ···················· | ····················· | ···················· | ···················· | ···················· |
| T | 4726 | hCoV-19/England/QEUH-B7C2D1/2020\|EPI_ISL_668161\|20 | ···················· | ···················· | ····················· | ···················· | ···················· | ···················· |
| T | 4727 | hCoV-19/England/QEUH-B820CF/2020\|EPI_ISL_668162\|20 | ···················· | ···················· | ····················· | ···················· | ···················· | ···················· |
| T | 4728 | hCoV-19/England/QEUH-B80674/2020\|EPI_ISL_668165\|20 | ···················· | ···················· | ····················· | ···················· | ···················· | ···················· |
| T | 4729 | hCoV-19/England/QEUH-B80717/2020\|EPI_ISL_668167\|20 | ···················· | ·A·················· | ····················· | ···················· | ···················· | ···A················ |
| T | 4730 | hCoV-19/England/QEUH-B80665/2020\|EPI_ISL_668168\|20 | ···················· | ···················· | ····················· | ···················· | ···················· | ···················· |
| T | 4731 | hCoV-19/England/CAMC-B776CB/2020\|EPI_ISL_668169\|20 | ···················· | ···················· | ····················· | ···················· | ······T············· | ···················· |
| T | 4732 | hCoV-19/England/CAMC-B776DA/2020\|EPI_ISL_668171\|20 | ···················· | ···················· | ····················· | ···················· | ······T············· | ···················· |
| T | 4733 | hCoV-19/England/QEUH-B8144C/2020\|EPI_ISL_668173\|20 | ···················· | ···················· | ····················· | ···················· | ···················· | ···················· |
| T | 4734 | hCoV-19/England/QEUH-B7C040/2020\|EPI_ISL_668174\|20 | ···················· | ·A·················· | ····················· | ···················· | ···················· | ···················· |
| T | 4735 | hCoV-19/England/QEUH-B7BFAE/2020\|EPI_ISL_668176\|20 | ···················· | ···················· | ····················· | ···················· | ···················· | ···················· |
| T | 4736 | hCoV-19/England/QEUH-B7FD0A/2020\|EPI_ISL_668177\|20 | ···················· | ···················· | ····················· | ···················· | ···················· | ···················· |
| T | 4737 | hCoV-19/England/QEUH-B7F696/2020\|EPI_ISL_668178\|20 | ···················· | ···················· | ····················· | ···················· | ···················· | ···················· |
| T | 4738 | hCoV-19/England/CAMC-B77616/2020\|EPI_ISL_668180\|20 | ···················· | ···················· | ····················· | ···················· | ···················· | ···················· |
| T | 4739 | hCoV-19/England/QEUH-B7BFBD/2020\|EPI_ISL_668181\|20 | ···················· | ···················· | ····················· | ···················· | ···················· | ···················· |
| T | 4740 | hCoV-19/England/QEUH-B7F924/2020\|EPI_ISL_668182\|20 | ···················· | ···················· | ····················· | ···················· | ···················· | ···················· |
| T | 4741 | hCoV-19/England/QEUH-B7C16B/2020\|EPI_ISL_668183\|20 | ···················· | ···················· | ····················· | ···················· | ···················· | ···················· |
| T | 4742 | hCoV-19/England/QEUH-B7C295/2020\|EPI_ISL_668186\|20 | ···················· | ···················· | ····················· | ···················· | ···················· | ···················· |
| T | 4743 | hCoV-19/England/QEUH-B82108/2020\|EPI_ISL_668189\|20 | ···················· | ···················· | ····················· | ···················· | ···················· | ···················· |
| T | 4744 | hCoV-19/England/CAMC-B77722/2020\|EPI_ISL_668190\|20 | ···················· | ·A·················· | ····················· | ···················· | ···················· | ···A················ |
| T | 4745 | hCoV-19/England/CAMC-B7787A/2020\|EPI_ISL_668192\|20 | ···················· | ···················· | ····················· | ···················· | ···················· | ···················· |
| T | 4746 | hCoV-19/England/QEUH-B7FDA0/2020\|EPI_ISL_668193\|20 | ···················· | ···················· | ····················· | ···················· | ···················· | ···················· |
| T | 4747 | hCoV-19/England/QEUH-B821EA/2020\|EPI_ISL_668196\|20 | ···················· | ···················· | ····················· | ···················· | ···················· | ···················· |
| T | 4751 | hCoV-19/England/CAMC-B78114/2020\|EPI_ISL_668200\|20 | ···················· | ···················· | ····················· | ···················· | ···················· | ···················· |
| T | 4752 | hCoV-19/England/CAMC-B77449/2020\|EPI_ISL_668201\|20 | ···················· | ···················· | ····················· | ···················· | ···················· | ···················· |
| T | 4753 | hCoV-19/England/QEUH-B7E80A/2020\|EPI_ISL_668202\|20 | ···················· | ···················· | ····················· | ···················· | ···················· | ···················· |
| T | 4754 | hCoV-19/England/QEUH-B7C07D/2020\|EPI_ISL_668203\|20 | ···················· | ···················· | ····················· | ···················· | ···················· | ···················· |
| T | 4755 | hCoV-19/England/QEUH-B7E873/2020\|EPI_ISL_668205\|20 | ···················· | ···················· | ····················· | ···················· | ···················· | ···················· |
| T | 4756 | hCoV-19/England/QEUH-B7FDBF/2020\|EPI_ISL_668206\|20 | ···················· | ···················· | ····················· | ···················· | ···················· | ···················· |
| T | 4757 | hCoV-19/England/QEUH-B7C05F/2020\|EPI_ISL_668208\|20 | ···················· | ···················· | ····················· | ···················· | ···················· | ···················· |
| T | 4758 | hCoV-19/England/QEUH-B7E8BF/2020\|EPI_ISL_668209\|20 | ···················· | ···················· | ····················· | ···················· | ···················· | ···················· |
| T | 4759 | hCoV-19/England/CAMC-B77889/2020\|EPI_ISL_668212\|20 | ···················· | ···················· | ····················· | ···················· | ···················· | ···················· |
| T | 4760 | hCoV-19/England/QEUH-B824F0/2020\|EPI_ISL_668215\|20 | ···················· | ···················· | ····················· | ···················· | ···················· | ···················· |
| T | 4762 | hCoV-19/England/QEUH-B7C444/2020\|EPI_ISL_668217\|20 | ···················· | ·A·················· | ····················· | ···················· | ···················· | ···················· |
| T | 4763 | hCoV-19/England/QEUH-B7C2FF/2020\|EPI_ISL_668218\|20 | ···················· | ···················· | ····················· | ···················· | ···················· | ···················· |
| T | 4764 | hCoV-19/England/QEUH-B7E855/2020\|EPI_ISL_668220\|20 | ···················· | ···················· | ····················· | ···················· | ···················· | ···················· |
| T | 4765 | hCoV-19/England/CAMC-B776AD/2020\|EPI_ISL_668222\|20 | ···················· | ···················· | ····················· | ···················· | ···················· | ···················· |
| T | 4766 | hCoV-19/England/QEUH-B7C013/2020\|EPI_ISL_668223\|20 | ···················· | ···················· | ····················· | ···················· | ···················· | ···················· |
| T | 4767 | hCoV-19/England/QEUH-B7C347/2020\|EPI_ISL_668224\|20 | ···················· | ···················· | ····················· | ···················· | ···················· | ···················· |
| T | 4768 | hCoV-19/England/QEUH-B7C022/2020\|EPI_ISL_668226\|20 | ···················· | ···················· | ····················· | ···················· | ···················· | ···················· |
| T | 4769 | hCoV-19/England/QEUH-B7C004/2020\|EPI_ISL_668229\|20 | ···················· | ·A·················· | ····················· | ···················· | ···················· | ···················· |
| T | 4770 | hCoV-19/England/QEUH-B823A8/2020\|EPI_ISL_668230\|20 | ···················· | ···················· | ····················· | ···················· | ···················· | ···················· |
| T | 4771 | hCoV-19/England/QEUH-B814C4/2020\|EPI_ISL_668231\|20 | ···················· | ···················· | ····················· | ···················· | ···················· | ···················· |
| T | 4772 | hCoV-19/England/QEUH-B7E460/2020\|EPI_ISL_668235\|20 | ···················· | ···················· | ····················· | ···················· | ···················· | ···················· |
| T | 4773 | hCoV-19/England/QEUH-B814B5/2020\|EPI_ISL_668236\|20 | ···················· | ···················· | ····················· | ···················· | ···················· | ···················· |
| T | 4774 | hCoV-19/Denmark/DCGC-10203/2020\|EPI_ISL_669450\|202 | ···················· | ···················· | ····················· | ···················· | ···················· | ···················· |
| T | 4775 | hCoV-19/Denmark/DCGC-10204/2020\|EPI_ISL_669451\|202 | ···················· | ···················· | ····················· | ···················· | ···················· | ···················· |
| T | 4776 | hCoV-19/Denmark/DCGC-10205/2020\|EPI_ISL_669452\|202 | ···················· | ···················· | ····················· | ···················· | ···················· | ···················· |
| T | 4777 | hCoV-19/Denmark/DCGC-10206/2020\|EPI_ISL_669453\|202 | ···················· | ···················· | ····················· | ···················· | ···················· | ···················· |
| T | 4778 | hCoV-19/Denmark/DCGC-10207/2020\|EPI_ISL_669454\|202 | ···················· | ···················· | ····················· | ···················· | ···················· | ···················· |
| T | 4779 | hCoV-19/Denmark/DCGC-10208/2020\|EPI_ISL_669455\|202 | ···················· | ···················· | ····················· | ···················· | ···················· | ···················· |
| T | 4780 | hCoV-19/Denmark/DCGC-10209/2020\|EPI_ISL_669456\|202 | ···················· | ···················· | ····················· | ···················· | ···················· | ···················· |
| T | 4781 | hCoV-19/Denmark/DCGC-10210/2020\|EPI_ISL_669457\|202 | ···················· | ···················· | ····················· | ···················· | ···················· | ···················· |
| T | 4782 | hCoV-19/Denmark/DCGC-10289/2020\|EPI_ISL_669537\|202 | ···················· | ···················· | ····················· | ···················· | ···················· | ···················· |
| T | 4783 | hCoV-19/Denmark/DCGC-10324/2020\|EPI_ISL_669538\|202 | ···················· | ···················· | ····················· | ···················· | ···················· | ···················· |

T 4784 hCoV-19/Denmark/DCGC-10554/2020|EPI_ISL_669539|202 .................... .................... ...........N........ .................... .................... ....................
T 4785 hCoV-19/Denmark/DCGC-10776/2020|EPI_ISL_669540|202 .................... .................... .................... .................... .................... ....................
T 4786 hCoV-19/Denmark/DCGC-10918/2020|EPI_ISL_669541|202 .................... .................... .................... .................... .................... ....................
T 4787 hCoV-19/Denmark/DCGC-11046/2020|EPI_ISL_669542|202 .................... .................... .................... .................... .................... ....................
T 4788 hCoV-19/Denmark/DCGC-11075/2020|EPI_ISL_669543|202 .................... .................... .................... .................... .................... ....................
T 4789 hCoV-19/Denmark/DCGC-10778/2020|EPI_ISL_669544|202 .................... .................... .................... .................... .................... ....................
T 4790 hCoV-19/Denmark/DCGC-10846/2020|EPI_ISL_669545|202 .................... .................... .................... .................... .................... ....................
T 4791 hCoV-19/Denmark/DCGC-10544/2020|EPI_ISL_669546|202 .................... .................... .................... .................... .................... ....................
T 4792 hCoV-19/Denmark/DCGC-10367/2020|EPI_ISL_669549|202 .................... .................... .................... .................... .................... ....................
T 4793 hCoV-19/Denmark/DCGC-10497/2020|EPI_ISL_669550|202 .................... .................... .................... .................... .................... ....................
T 4794 hCoV-19/Denmark/DCGC-10614/2020|EPI_ISL_669551|202 .................... .................... .................... .................... .................... ....................
T 4795 hCoV-19/Denmark/DCGC-10811/2020|EPI_ISL_669552|202 .................... .................... .................... .................... .................... ....................
T 4796 hCoV-19/Denmark/DCGC-11098/2020|EPI_ISL_669553|202 .................... .................... .................... .................... .................... ....................
T 4797 hCoV-19/Denmark/DCGC-11138/2020|EPI_ISL_669554|202 .................... .................... .................... .................... .................... ....................
T 4798 hCoV-19/Denmark/DCGC-11189/2020|EPI_ISL_669555|202 .................... .................... .................... .................... .................... ....................
T 4799 hCoV-19/Denmark/DCGC-11208/2020|EPI_ISL_669556|202 .................... .................... .................... .................... .................... ....................
T 4800 hCoV-19/Denmark/DCGC-11166/2020|EPI_ISL_670430|202 .................... .A.................. .................... .................... .................... ....................
T 4801 hCoV-19/Denmark/DCGC-11167/2020|EPI_ISL_670431|202 .................... .................... .................... .................... .................... ....................
T 4802 hCoV-19/Denmark/DCGC-11168/2020|EPI_ISL_670432|202 .................... .................... .................... .................... .................... ....................
T 4803 hCoV-19/Denmark/DCGC-11169/2020|EPI_ISL_670433|202 .................... .................... .................... .................... .................... ....................
T 4804 hCoV-19/Denmark/DCGC-11170/2020|EPI_ISL_670434|202 .................... .................... .................... .................... .................... ....................
T 4805 hCoV-19/Denmark/DCGC-11171/2020|EPI_ISL_670435|202 .................... .................... .................... .................... .................... ....................
T 4806 hCoV-19/Denmark/DCGC-11172/2020|EPI_ISL_670436|202 .................... .................... .................... .................... .................... ....................
T 4807 hCoV-19/Denmark/DCGC-11173/2020|EPI_ISL_670437|202 .................... .................... .................... .................... .................... ....................
T 4808 hCoV-19/Denmark/DCGC-11175/2020|EPI_ISL_670438|202 .................... .A.................. .................... .................... .................... ...A................
T 4809 hCoV-19/Denmark/DCGC-11181/2020|EPI_ISL_670439|202 .................... .................... .................... .................... .................... ....................
T 4810 hCoV-19/Denmark/DCGC-11183/2020|EPI_ISL_670440|202 .................... .A.................. .................... .................... .................... ....................
T 4811 hCoV-19/Denmark/DCGC-11184/2020|EPI_ISL_670441|202 .................... .................... .................... .................... ......T............. ....................
T 4812 hCoV-19/Denmark/DCGC-11186/2020|EPI_ISL_670442|202 .................... .................... .................... .................... .................... ....................
T 4813 hCoV-19/Denmark/DCGC-11188/2020|EPI_ISL_670443|202 .................... .A.................. .................... .................... .................... ....................
T 4814 hCoV-19/Denmark/DCGC-11190/2020|EPI_ISL_670444|202 .................... .................... .................... .................... .................... ....................
T 4815 hCoV-19/Denmark/DCGC-11191/2020|EPI_ISL_670445|202 .................... .................... .................... .................... .................... ....................
T 4816 hCoV-19/Denmark/DCGC-11192/2020|EPI_ISL_670446|202 .................... .................... .................... .................... .................... ....................
T 4817 hCoV-19/Denmark/DCGC-11193/2020|EPI_ISL_670447|202 .................... .................... .................... .................... .................... ....................
T 4818 hCoV-19/Denmark/DCGC-11194/2020|EPI_ISL_670448|202 .................... .................... .................... .................... .................... ....................
T 4819 hCoV-19/Denmark/DCGC-11196/2020|EPI_ISL_670449|202 .................... .................... .................... .................... .................... ....................
T 4820 hCoV-19/Denmark/DCGC-11197/2020|EPI_ISL_670450|202 .................... .................... .................... .................... .................... ....................
T 4821 hCoV-19/Denmark/DCGC-11199/2020|EPI_ISL_670451|202 .................... .................... .................... .................... .................... ....................
T 4822 hCoV-19/Denmark/DCGC-11200/2020|EPI_ISL_670452|202 .................... .................... .................... .................... .................... ....................
T 4823 hCoV-19/Denmark/DCGC-11202/2020|EPI_ISL_670453|202 .................... .................... .................... .................... .................... ....................
T 4824 hCoV-19/Denmark/DCGC-11204/2020|EPI_ISL_670454|202 .................... .................... .................... .................... .................... ....................
T 4825 hCoV-19/Denmark/DCGC-11205/2020|EPI_ISL_670455|202 .................... .................... .................... .................... .................... ....................
T 4826 hCoV-19/Denmark/DCGC-11209/2020|EPI_ISL_670456|202 .................... .................... .................... .................... .................... ....................
T 4827 hCoV-19/Denmark/DCGC-11210/2020|EPI_ISL_670457|202 .................... .................... .................... .................... .................... ....................
T 4828 hCoV-19/Denmark/DCGC-11433/2020|EPI_ISL_670523|202 .................... .................... .................... .................... .................... ....................
T 4829 hCoV-19/Denmark/DCGC-11874/2020|EPI_ISL_670524|202 .................... .................... .................... .................... .................... ....................
T 4830 hCoV-19/Denmark/DCGC-11893/2020|EPI_ISL_670525|202 .................... .................... .................... .................... .................... ....................
T 4831 hCoV-19/Denmark/DCGC-11908/2020|EPI_ISL_670526|202 .................... .................... .................... .................... .................... ....................
T 4832 hCoV-19/Denmark/DCGC-11915/2020|EPI_ISL_670527|202 .................... .................... .................... .................... .................... ....................
T 4833 hCoV-19/Denmark/DCGC-11526/2020|EPI_ISL_670529|202 .................... .................... .................... .................... .................... ....................
T 4834 hCoV-19/Denmark/DCGC-11798/2020|EPI_ISL_670530|202 .................... .................... .................... .................... .................... ....................
T 4835 hCoV-19/Denmark/DCGC-11705/2020|EPI_ISL_670531|202 .................... .................... .................... .................... .................... ....................
T 4836 hCoV-19/USA/NY-NYCPHL-001183/2020|EPI_ISL_671762|2 .................... .................... .................... .................... .................... ....................
T 4838 hCoV-19/USA/CA-SCCPHD-UC200/2020|EPI_ISL_671768|20 .................... .................... .................... .................... .................... ....................
T 4847 hCoV-19/Ireland/CE-NVRL-75IRL33549/2020|EPI_ISL_67 .................... .................... .................... .................... .................... ....................
T 4848 hCoV-19/Ireland/CN-NVRL-75IRL94334/2020|EPI_ISL_67 .................... .................... .................... .................... .................... ....................
T 4849 hCoV-19/Ireland/CO-NVRL-20IRL79223/2020|EPI_ISL_67 .................... .................... .................... .................... .................... ....................
T 4850 hCoV-19/Ireland/CO-NVRL-20IRL79224/2020|EPI_ISL_67 .................... .................... .................... .................... .................... ....................
T 4851 hCoV-19/Ireland/CO-NVRL-75IRL23243/2020|EPI_ISL_67 .................... .................... .................... .................... .................... ....................
T 4852 hCoV-19/Ireland/CO-NVRL-75IRL23254/2020|EPI_ISL_67 .................... .................... .................... .................... .................... ....................
T 4853 hCoV-19/Ireland/CO-NVRL-75IRL24656/2020|EPI_ISL_67 .................... .................... .................... .................... .................... ....................
T 4854 hCoV-19/Ireland/CO-NVRL-75IRL24658/2020|EPI_ISL_67 .................... .................... .................... .................... .................... ....................
T 4855 hCoV-19/Ireland/CO-NVRL-75IRL32904/2020|EPI_ISL_67 .................... .................... .................... .................... .................... ....................
T 4856 hCoV-19/Ireland/CO-NVRL-75IRL33101/2020|EPI_ISL_67 .................... .................... .................... .................... .................... ....................
T 4857 hCoV-19/Ireland/CO-NVRL-75IRL33266/2020|EPI_ISL_67 .................... .................... .................... .................... .................... ....................
T 4858 hCoV-19/Ireland/CO-NVRL-75IRL33272/2020|EPI_ISL_67 .................... .................... .................... .................... .................... ....................
T 4859 hCoV-19/Ireland/CO-NVRL-75IRL33290/2020|EPI_ISL_67 .................... .................... .................... .................... .................... ....................
T 4860 hCoV-19/Ireland/CO-NVRL-75IRL33312/2020|EPI_ISL_67 .................... .................... .................... .................... .................... ....................
T 4861 hCoV-19/Ireland/CO-NVRL-75IRL73629/2020|EPI_ISL_67 .................... .................... .................... .................... .................... ....................
T 4862 hCoV-19/Ireland/CO-NVRL-75IRL73676/2020|EPI_ISL_67 .................... .................... .................... .................... .................... ....................
T 4863 hCoV-19/Ireland/CO-NVRL-75IRL75321/2020|EPI_ISL_67 .................... .................... .................... .................... .................... ....................
T 4864 hCoV-19/Ireland/CO-NVRL-75IRL84314/2020|EPI_ISL_67 .................... .................... .................... .................... .................... ....................
T 4865 hCoV-19/England/ALDP-B7478F/2020|EPI_ISL_662000|20 .................... .................... .................... .................... .................... ....................
T 4866 hCoV-19/England/ALDP-B74655/2020|EPI_ISL_662001|20 .................... .................... .................... .................... .................... ....................
T 4867 hCoV-19/England/ALDP-B749F2/2020|EPI_ISL_662002|20 .................... .................... .................... .................... .................... ....................
T 4868 hCoV-19/England/QEUH-B68D8C/2020|EPI_ISL_662003|20 .................... .................... .................... .................... .................... ....................
T 4869 hCoV-19/England/ALDP-B75593/2020|EPI_ISL_662004|20 .................... .................... .................... .................... .................... ....................
T 4870 hCoV-19/England/QEUH-B68153/2020|EPI_ISL_662005|20 .................... .................... .................... .................... .................... ....................
T 4871 hCoV-19/England/ALDP-B757D9/2020|EPI_ISL_662006|20 .................... .................... .................... .................... .................... ....................
T 4872 hCoV-19/England/ALDP-B75618/2020|EPI_ISL_662008|20 .................... .................... .................... .................... .................... ....................

| | | | | | | | | |
|---|---|---|---|---|---|---|---|---|
| T | 4873 hCoV-19/England/QEUH-B6741F/2020\|EPI_ISL_662009\|20 | ···················· | ···················· | ····················· | ···················· | ···················· | ···················· | |
| T | 4874 hCoV-19/England/ALDP-B740A2/2020\|EPI_ISL_662010\|20 | ···················· | ···················· | ····················· | ···················· | ···················· | ···················· | |
| T | 4875 hCoV-19/England/QEUH-B68083/2020\|EPI_ISL_662011\|20 | ···················· | ···················· | ····················· | ···················· | ···················· | ···················· | |
| T | 4876 hCoV-19/England/QEUH-B6918F/2020\|EPI_ISL_662012\|20 | ···················· | ···················· | ····················· | ···················· | ···················· | ···················· | |
| T | 4877 hCoV-19/England/ALDP-B7439A/2020\|EPI_ISL_662013\|20 | ···················· | ···················· | ····················· | ···················· | ···················· | ···················· | |
| T | 4878 hCoV-19/England/ALDP-B74673/2020\|EPI_ISL_662014\|20 | ···················· | ···················· | ····················· | ···················· | ···················· | ···················· | |
| T | 4879 hCoV-19/France/NAQ-SC286/2020\|EPI_ISL_663225\|2020- | ···················· | ···················· | ····················· | ···················· | ···················· | ···················· | |
| T | 4880 hCoV-19/France/NAQ-SC287/2020\|EPI_ISL_663226\|2020- | ···················· | ···················· | ····················· | ···················· | ···················· | ···················· | |
| T | 4881 hCoV-19/France/NAQ-SC576/2020\|EPI_ISL_663227\|2020- | ···················· | ···················· | ····················· | ···················· | ···················· | ···················· | |
| T | 4882 hCoV-19/France/NAQ-SC577/2020\|EPI_ISL_663228\|2020- | ···················· | ···················· | ····················· | ···················· | NNN················· | ···················· | 1 |
| T | 4883 hCoV-19/France/NAQ-SC578/2020\|EPI_ISL_663229\|2020- | ···················· | ···················· | ····················· | ···················· | ···················· | ···················· | |
| T | 4884 hCoV-19/France/NAQ-SC579/2020\|EPI_ISL_663230\|2020- | ···················· | ···················· | ····················· | ···················· | ···················· | ···················· | |
| T | 4885 hCoV-19/France/NAQ-SC580/2020\|EPI_ISL_663231\|2020- | ···················· | ···················· | ····················· | ···················· | ···················· | ···················· | |
| T | 4887 hCoV-19/France/NAQ-SC582/2020\|EPI_ISL_663233\|2020- | ···················· | ···················· | ····················· | ···················· | ···················· | ···················· | |
| T | 4888 hCoV-19/France/NAQ-SC583/2020\|EPI_ISL_663234\|2020- | ···················· | ···················· | ····················· | ···················· | ···················· | ···················· | |
| T | 4889 hCoV-19/France/NAQ-SC584/2020\|EPI_ISL_663235\|2020- | ···················· | ···················· | ····················· | ···················· | ···················· | ···················· | |
| T | 4890 hCoV-19/France/NAQ-SC585/2020\|EPI_ISL_663236\|2020- | ···················· | ···················· | ····················· | ···················· | ···················· | ···················· | |
| T | 4891 hCoV-19/France/NAQ-SC587/2020\|EPI_ISL_663237\|2020- | ···················· | ···················· | ····················· | ···················· | ···················· | ···················· | |
| T | 4892 hCoV-19/France/NAQ-SC592/2020\|EPI_ISL_663239\|2020- | ···················· | ···················· | ····················· | ···················· | ···················· | ···················· | |
| T | 4893 hCoV-19/France/NAQ-SC596/2020\|EPI_ISL_663240\|2020- | ···················· | ···················· | ····················· | ···················· | ···················· | ···················· | |
| T | 4894 hCoV-19/France/NAQ-SC597/2020\|EPI_ISL_663241\|2020- | ···················· | ···················· | ····················· | ···················· | ···················· | ···················· | |
| T | 4896 hCoV-19/France/OCC-SC566/2020\|EPI_ISL_663243\|2020- | ···················· | ··········A········· | ····················· | ···················· | ···················· | ···················· | |
| T | 4897 hCoV-19/France/OCC-SC567/2020\|EPI_ISL_663244\|2020- | ···················· | ···················· | ····················· | ···················· | ···················· | ···················· | |
| T | 4898 hCoV-19/France/OCC-SC568/2020\|EPI_ISL_663245\|2020- | ···················· | ···················· | ····················· | ···················· | ···················· | ···················· | |
| T | 4899 hCoV-19/France/OCC-SC569/2020\|EPI_ISL_663246\|2020- | ···················· | ···················· | ····················· | ···················· | ···················· | ···················· | |
| T | 4900 hCoV-19/France/OCC-SC570/2020\|EPI_ISL_663247\|2020- | ···················· | ···················· | ····················· | ···················· | ···················· | ···················· | |
| T | 4901 hCoV-19/France/OCC-SC571/2020\|EPI_ISL_663248\|2020- | ···················· | ···················· | ····················· | ···················· | ···················· | ···················· | |
| T | 4902 hCoV-19/France/PDL-SC599/2020\|EPI_ISL_663249\|2020- | ···················· | ···················· | ····················· | ···················· | ··N················· | ···················· | |
| T | 4903 hCoV-19/France/PDL-SC602/2020\|EPI_ISL_663250\|2020- | ···················· | ···················· | ····················· | ···················· | ···················· | ···················· | |
| T | 4904 hCoV-19/France/PDL-SC603/2020\|EPI_ISL_663251\|2020- | ···················· | ···················· | ····················· | ···················· | ···················· | ···················· | |
| T | 4905 hCoV-19/France/PDL-SC604/2020\|EPI_ISL_663252\|2020- | ···················· | ···················· | ····················· | ···················· | ···················· | ···················· | |
| T | 4906 hCoV-19/France/PDL-SC605/2020\|EPI_ISL_663253\|2020- | ···················· | ···················· | ····················· | ···················· | ···················· | ···················· | |
| T | 4907 hCoV-19/France/PDL-SC607/2020\|EPI_ISL_663254\|2020- | ···················· | ···················· | ····················· | ···················· | ···················· | ···················· | |
| T | 4908 hCoV-19/France/PDL-SC608/2020\|EPI_ISL_663255\|2020- | ···················· | ···················· | ····················· | ···················· | ···················· | ···················· | |
| T | 4909 hCoV-19/France/PDL-SC609/2020\|EPI_ISL_663256\|2020- | ···················· | ···················· | ····················· | ···················· | ···················· | ···················· | |
| T | 4910 hCoV-19/France/PDL-SC611/2020\|EPI_ISL_663257\|2020- | ···················· | ···················· | ····················· | ···················· | ···················· | ···················· | |
| T | 4911 hCoV-19/France/PDL-SC612/2020\|EPI_ISL_663258\|2020- | ···················· | ···················· | ····················· | ···················· | ···················· | ···················· | |
| T | 4913 hCoV-19/France/PDL-SC614/2020\|EPI_ISL_663260\|2020- | ···················· | ···················· | ····················· | ···················· | N··················· | ···················· | |
| T | 4914 hCoV-19/France/PDL-SC615/2020\|EPI_ISL_663261\|2020- | ···················· | ···················· | ····················· | ···················· | ···················· | ···················· | |
| T | 4915 hCoV-19/France/PDL-SC616/2020\|EPI_ISL_663262\|2020- | ···················· | ···················· | ····················· | ···················· | ···················· | ···················· | |
| T | 4916 hCoV-19/France/PDL-SC617/2020\|EPI_ISL_663263\|2020- | ···················· | ···················· | ····················· | ···················· | ···················· | ···················· | |
| T | 4917 hCoV-19/France/PDL-SC620/2020\|EPI_ISL_663264\|2020- | ···················· | ···················· | ····················· | ···················· | ···················· | ···················· | |
| T | 4919 hCoV-19/France/PDL-SC623/2020\|EPI_ISL_663266\|2020- | ···················· | ···················· | ····················· | ···················· | ···················· | ···················· | |
| T | 4920 hCoV-19/France/PDL-SC624/2020\|EPI_ISL_663267\|2020- | ···················· | ···················· | ····················· | ···················· | ···················· | ···················· | |
| T | 4921 hCoV-19/France/PDL-SC625/2020\|EPI_ISL_663268\|2020- | ···················· | ···················· | ····················· | ···················· | ···················· | ···················· | |
| T | 4922 hCoV-19/France/PDL-SC626/2020\|EPI_ISL_663269\|2020- | ···················· | ···················· | ····················· | ···················· | NNN················· | ···················· | 1 |
| T | 4923 hCoV-19/France/PDL-SC629/2020\|EPI_ISL_663270\|2020- | ···················· | ···················· | ····················· | ···················· | ···················· | ···················· | |
| T | 4924 hCoV-19/France/PDL-SC630/2020\|EPI_ISL_663271\|2020- | ···················· | ···················· | ····················· | ···················· | ···················· | ···················· | |
| T | 4925 hCoV-19/France/PDL-SC632/2020\|EPI_ISL_663272\|2020- | ···················· | ···················· | ····················· | ···················· | ···················· | ···················· | |
| T | 4926 hCoV-19/France/PDL-SC634/2020\|EPI_ISL_663273\|2020- | ···················· | ···················· | ····················· | ···················· | ···················· | ···················· | |
| T | 4927 hCoV-19/France/PDL-SC635/2020\|EPI_ISL_663274\|2020- | ···················· | ···················· | ····················· | ···················· | ···················· | ···················· | |
| T | 4928 hCoV-19/France/PDL-SC636/2020\|EPI_ISL_663275\|2020- | ···················· | ···················· | ····················· | ···················· | ···················· | ···················· | |
| T | 4929 hCoV-19/France/PDL-SC640/2020\|EPI_ISL_663278\|2020- | ···················· | ···················· | ····················· | ···················· | ···················· | ···················· | |
| T | 4930 hCoV-19/France/PDL-SC641/2020\|EPI_ISL_663279\|2020- | ···················· | ···················· | ····················· | ···················· | ···················· | ···················· | |
| T | 4931 hCoV-19/France/PDL-SC642/2020\|EPI_ISL_663280\|2020- | ···················· | ···················· | ····················· | ···················· | ···················· | ···················· | |
| T | 4932 hCoV-19/France/PDL-SC643/2020\|EPI_ISL_663281\|2020- | ···················· | ···················· | ····················· | ···················· | ···················· | ···················· | |
| T | 4933 hCoV-19/France/PDL-SC644/2020\|EPI_ISL_663282\|2020- | ···················· | ···················· | ····················· | ···················· | ···················· | ···················· | |
| T | 4934 hCoV-19/France/PDL-SC646/2020\|EPI_ISL_663283\|2020- | ···················· | ···················· | ····················· | ···················· | ···················· | ···················· | |
| T | 4935 hCoV-19/France/PDL-SC647/2020\|EPI_ISL_663284\|2020- | ···················· | ···················· | ····················· | ···················· | ···················· | ···················· | |
| T | 4937 hCoV-19/England/CAMC-B2345B/2020\|EPI_ISL_664604\|20 | ···················· | ·A·················· | ····················· | ···················· | ···················· | ··A················· | |
| T | 4944 hCoV-19/England/NORW-F57CD/2020\|EPI_ISL_665893\|202 | ···················· | ···················· | ····················· | ···················· | ···················· | ···················· | |
| T | 4949 hCoV-19/England/NOTT-11A3AF/2020\|EPI_ISL_665898\|20 | ···················· | ···················· | ····················· | ···················· | ···················· | ···················· | |
| T | 4950 hCoV-19/England/NOTT-11B0E3/2020\|EPI_ISL_665899\|20 | ···················· | ···················· | ····················· | ···················· | ···················· | ···················· | |
| T | 4951 hCoV-19/England/NOTT-11D236/2020\|EPI_ISL_665920\|20 | ···················· | ···················· | ····················· | ···················· | ···················· | ···················· | |
| T | 4952 hCoV-19/England/NOTT-11D245/2020\|EPI_ISL_665921\|20 | ···················· | ···················· | ····················· | ···················· | ···················· | ···················· | |
| T | 4953 hCoV-19/England/NOTT-11D263/2020\|EPI_ISL_665922\|20 | ···················· | ···················· | ····················· | ···················· | ···················· | ···················· | |
| T | 4954 hCoV-19/England/NOTT-11D272/2020\|EPI_ISL_665923\|20 | ···················· | ···················· | ····················· | ···················· | ···················· | ···················· | |
| T | 4955 hCoV-19/England/NOTT-11D281/2020\|EPI_ISL_665924\|20 | ···················· | ···················· | ····················· | ···················· | ···················· | ···················· | |
| T | 4956 hCoV-19/England/NOTT-11D315/2020\|EPI_ISL_665925\|20 | ···················· | ···················· | ····················· | ···················· | ···················· | ···················· | |
| T | 4957 hCoV-19/England/NOTT-11D333/2020\|EPI_ISL_665926\|20 | ···················· | ···················· | ····················· | ···················· | ···················· | ···················· | |
| T | 4958 hCoV-19/England/NOTT-11D3BB/2020\|EPI_ISL_665927\|20 | ···················· | ···················· | ····················· | ···················· | ···················· | ···················· | |
| T | 4959 hCoV-19/England/NOTT-11D3CA/2020\|EPI_ISL_665928\|20 | ···················· | ···················· | ····················· | ···················· | ···················· | ···················· | |
| T | 4960 hCoV-19/England/NOTT-11D3D9/2020\|EPI_ISL_665929\|20 | ···················· | ···················· | ····················· | ···················· | ···················· | ···················· | |
| T | 4961 hCoV-19/England/NOTT-11D48B/2020\|EPI_ISL_665930\|20 | ···················· | ···················· | ····················· | ···················· | ···················· | ···················· | |
| T | 4962 hCoV-19/England/NOTT-11D49A/2020\|EPI_ISL_665931\|20 | ···················· | ···················· | ····················· | ···················· | ···················· | ···················· | |
| T | 4963 hCoV-19/England/NOTT-11D4A9/2020\|EPI_ISL_665932\|20 | ···················· | ···················· | ····················· | ···················· | ···················· | ···················· | |
| T | 4964 hCoV-19/England/NOTT-11D4B8/2020\|EPI_ISL_665933\|20 | ···················· | ···················· | ····················· | ···················· | ···················· | ···················· | |
| T | 4965 hCoV-19/England/NOTT-11D4D6/2020\|EPI_ISL_665934\|20 | ···················· | ···················· | ····················· | ···················· | ···················· | ···················· | |
| T | 4966 hCoV-19/England/NOTT-11D53D/2020\|EPI_ISL_665935\|20 | ···················· | ···················· | ····················· | ···················· | ···················· | ···················· | |
| T | 4967 hCoV-19/England/NOTT-11D56A/2020\|EPI_ISL_665936\|20 | ···················· | ···················· | ····················· | ···················· | ···················· | ···················· | |

| | | | | | | | |
|---|---|---|---|---|---|---|---|
| T | 4968 hCoV-19/England/NOTT-11D5A6/2020\|EPI_ISL_665937\|20 | ···················· | ···················· | ···················· | ···················· | ···················· | ···················· |
| T | 4969 hCoV-19/England/NOTT-11D685/2020\|EPI_ISL_665938\|20 | ···················· | ···················· | ···················· | ···················· | ···················· | ···················· |
| T | 4970 hCoV-19/England/NOTT-11D755/2020\|EPI_ISL_665939\|20 | ···················· | ···················· | ···················· | ···················· | ···················· | ···················· |
| T | 4971 hCoV-19/England/NOTT-11D7FB/2020\|EPI_ISL_665940\|20 | ···················· | ···················· | ···················· | ···················· | ···················· | ···················· |
| T | 4972 hCoV-19/England/NOTT-11D807/2020\|EPI_ISL_665941\|20 | ···················· | ···················· | ···················· | ···················· | ···················· | ···················· |
| T | 4973 hCoV-19/England/NOTT-11D852/2020\|EPI_ISL_665942\|20 | ···················· | ·A·················· | ···················· | ···················· | ···················· | ··A················· |
| T | 4974 hCoV-19/England/NOTT-11D88F/2020\|EPI_ISL_665943\|20 | ···················· | ···················· | ···················· | ···················· | ···················· | ···················· |
| T | 4975 hCoV-19/England/NOTT-11D8BC/2020\|EPI_ISL_665944\|20 | ···················· | ···················· | ···················· | ···················· | ···················· | ···················· |
| T | 4976 hCoV-19/England/NOTT-11D8DA/2020\|EPI_ISL_665945\|20 | ···················· | ···················· | ···················· | ···················· | ···················· | ···················· |
| T | 4978 hCoV-19/England/PORT-2EA1BC/2020\|EPI_ISL_665947\|20 | ···················· | ···················· | ···················· | ···················· | ···················· | ···················· |
| T | 4979 hCoV-19/England/PORT-2EA1F8/2020\|EPI_ISL_665948\|20 | ···················· | ···················· | ···················· | ···················· | ···················· | ···················· |
| T | 4980 hCoV-19/England/PORT-2EA25F/2020\|EPI_ISL_665949\|20 | ···················· | ···················· | ···················· | ···················· | ······T············· | ···················· |
| T | 4981 hCoV-19/England/PORT-2EA2AA/2020\|EPI_ISL_665950\|20 | ···················· | ···················· | ···················· | ···················· | ······T············· | ···················· |
| T | 4982 hCoV-19/England/PORT-2EA2C8/2020\|EPI_ISL_665951\|20 | ···················· | ···················· | ···················· | ···················· | ···················· | ···················· |
| T | 4983 hCoV-19/England/PORT-2EA3B6/2020\|EPI_ISL_665952\|20 | ···················· | ···················· | ···················· | ···················· | ···················· | ···················· |
| T | 4984 hCoV-19/England/PORT-2EA3D4/2020\|EPI_ISL_665953\|20 | ···················· | ···················· | ···················· | ···················· | ···················· | ···················· |
| T | 4985 hCoV-19/England/PORT-2EA495/2020\|EPI_ISL_665954\|20 | ···················· | ···················· | ···················· | ···················· | ···················· | ···················· |
| T | 4986 hCoV-19/England/PORT-2EA50B/2020\|EPI_ISL_665955\|20 | ···················· | ···················· | ···················· | ···················· | ······T············· | ···················· |
| T | 4987 hCoV-19/England/QEUH-9AE566/2020\|EPI_ISL_665956\|20 | ···················· | ···················· | ···················· | ···················· | ···················· | ···················· |
| T | 4988 hCoV-19/England/QEUH-A313F4/2020\|EPI_ISL_665957\|20 | ···················· | ···················· | ···················· | ···················· | ···················· | ···················· |
| T | 4989 hCoV-19/England/QEUH-A31567/2020\|EPI_ISL_665958\|20 | ···················· | ···················· | ···················· | ···················· | ···················· | ···················· |
| T | 4990 hCoV-19/England/QEUH-A490A7/2020\|EPI_ISL_665959\|20 | ···················· | ···················· | ···················· | ···················· | ···················· | ···················· |
| T | 4991 hCoV-19/England/QEUH-A493CC/2020\|EPI_ISL_665960\|20 | ···················· | ···················· | ···················· | ···················· | ····T··············· | ···················· |
| T | 4992 hCoV-19/England/QEUH-A4CE6E/2020\|EPI_ISL_665961\|20 | ···················· | ···················· | ···················· | ···················· | ······T············· | ···················· |
| T | 4993 hCoV-19/England/QEUH-A4D049/2020\|EPI_ISL_665962\|20 | ···················· | ···················· | ···················· | ···················· | ···················· | ···················· |
| T | 4994 hCoV-19/England/QEUH-A4D0EF/2020\|EPI_ISL_665963\|20 | ···················· | ···················· | ···················· | ···················· | ···················· | ···················· |
| T | 4995 hCoV-19/England/QEUH-A60505/2020\|EPI_ISL_665964\|20 | ···················· | ···················· | ···················· | ···················· | ···················· | ···················· |
| T | 4996 hCoV-19/England/QEUH-A605E7/2020\|EPI_ISL_665965\|20 | ···················· | ···················· | ···················· | ···················· | ···················· | ···················· |
| T | 4997 hCoV-19/England/QEUH-A60B03/2020\|EPI_ISL_665966\|20 | ···················· | ···················· | ···················· | ···················· | ···················· | ···················· |
| T | 4998 hCoV-19/England/QEUH-A60BC7/2020\|EPI_ISL_665967\|20 | ···················· | ···················· | ···················· | ···················· | ···················· | ···················· |
| T | 4999 hCoV-19/England/QEUH-A60CC4/2020\|EPI_ISL_665968\|20 | ···················· | ···················· | ···················· | ···················· | ···················· | ···················· |
| T | 5000 hCoV-19/England/QEUH-A60D0D/2020\|EPI_ISL_665969\|20 | ···················· | ···················· | ···················· | ···················· | ···················· | ···················· |
| T | 5001 hCoV-19/England/QEUH-A60D2B/2020\|EPI_ISL_665970\|20 | ···················· | ···················· | ···················· | ···················· | ···················· | ···················· |
| T | 5002 hCoV-19/England/QEUH-A7635D/2020\|EPI_ISL_665971\|20 | ···················· | ···················· | ···················· | ···················· | ···················· | ···················· |
| T | 5003 hCoV-19/England/QEUH-A7636C/2020\|EPI_ISL_665972\|20 | ···················· | ···················· | ···················· | ···················· | ···················· | ···················· |
| T | 5004 hCoV-19/England/QEUH-A76760/2020\|EPI_ISL_665973\|20 | ···················· | ···················· | ···················· | ···················· | ···················· | ···················· |
| T | 5005 hCoV-19/England/QEUH-A7678E/2020\|EPI_ISL_665974\|20 | ···················· | ···················· | ···················· | ···················· | ···················· | ···················· |
| T | 5006 hCoV-19/England/QEUH-A76BCE/2020\|EPI_ISL_665975\|20 | ···················· | ···················· | ···················· | ···················· | ···················· | ···················· |
| T | 5007 hCoV-19/England/QEUH-A76C8F/2020\|EPI_ISL_665976\|20 | ···················· | ···················· | ···················· | ···················· | ···················· | ···················· |
| T | 5008 hCoV-19/England/QEUH-A9BFC7/2020\|EPI_ISL_665977\|20 | ···················· | ···················· | ···················· | ···················· | ···················· | ···················· |
| T | 5009 hCoV-19/England/QEUH-A9BFE5/2020\|EPI_ISL_665978\|20 | ···················· | ·A·················· | ···················· | ···················· | ···················· | ··A················· |
| T | 5010 hCoV-19/England/QEUH-A9C8E9/2020\|EPI_ISL_665979\|20 | ···················· | ···················· | ···················· | ···················· | ···················· | ···················· |
| T | 5011 hCoV-19/England/QEUH-A9C95F/2020\|EPI_ISL_665980\|20 | ···················· | ···················· | ···················· | ···················· | ···················· | ···················· |
| T | 5012 hCoV-19/England/QEUH-A9C99B/2020\|EPI_ISL_665981\|20 | ···················· | ···················· | ···················· | ···················· | ···················· | ···················· |
| T | 5013 hCoV-19/England/QEUH-A9D1A1/2020\|EPI_ISL_665982\|20 | ···················· | ···················· | ···················· | ···················· | ···················· | ···················· |
| T | 5014 hCoV-19/England/QEUH-A9F5A3/2020\|EPI_ISL_665983\|20 | ···················· | ···················· | ···················· | ···················· | ···················· | ···················· |
| T | 5015 hCoV-19/England/QEUH-A9F78F/2020\|EPI_ISL_665984\|20 | ···················· | ···················· | ···················· | ···················· | ···················· | ···················· |
| T | 5016 hCoV-19/England/QEUH-A9F7AD/2020\|EPI_ISL_665985\|20 | ···················· | ···················· | ···················· | ···················· | ···················· | ···················· |
| T | 5017 hCoV-19/England/QEUH-A9F7BC/2020\|EPI_ISL_665986\|20 | ···················· | ···················· | ···················· | ···················· | ···················· | ···················· |
| T | 5018 hCoV-19/England/QEUH-A9FA1D/2020\|EPI_ISL_665987\|20 | ···················· | ···················· | ···················· | ···················· | ···················· | ···················· |
| T | 5019 hCoV-19/England/QEUH-A9FA3B/2020\|EPI_ISL_665988\|20 | ···················· | ···················· | ···················· | ···················· | ···················· | ···················· |
| T | 5020 hCoV-19/England/QEUH-A9FA77/2020\|EPI_ISL_665989\|20 | ···················· | ···················· | ···················· | ···················· | ······T············· | ···············A···· |
| T | 5021 hCoV-19/England/QEUH-A9FA86/2020\|EPI_ISL_665990\|20 | ···················· | ···················· | ···················· | ···················· | ···················· | ···················· |
| T | 5022 hCoV-19/England/QEUH-A9FAA4/2020\|EPI_ISL_665991\|20 | ···················· | ···················· | ···················· | ···················· | ···················· | ···················· |
| T | 5023 hCoV-19/England/QEUH-A9FC08/2020\|EPI_ISL_665992\|20 | ···················· | ···················· | ···················· | ···················· | ···················· | ···················· |
| T | 5024 hCoV-19/England/QEUH-A9FC26/2020\|EPI_ISL_665993\|20 | ···················· | ···················· | ···················· | ···················· | ···················· | ···················· |
| T | 5025 hCoV-19/England/QEUH-A9FC35/2020\|EPI_ISL_665994\|20 | ···················· | ···················· | ···················· | ···················· | ···················· | ···················· |
| T | 5026 hCoV-19/England/QEUH-A9FC71/2020\|EPI_ISL_665995\|20 | ···················· | ···················· | ···················· | ···················· | ···················· | ···················· |
| T | 5027 hCoV-19/England/QEUH-A9FCAE/2020\|EPI_ISL_665996\|20 | ···················· | ···················· | ···················· | ···················· | ···················· | ···················· |
| T | 5028 hCoV-19/England/QEUH-A9FF1E/2020\|EPI_ISL_665997\|20 | ···················· | ···················· | ···················· | ···················· | ···················· | ···················· |
| T | 5029 hCoV-19/England/QEUH-AA0448/2020\|EPI_ISL_665998\|20 | ···················· | ···················· | ···················· | ···················· | ···················· | ···················· |
| T | 5030 hCoV-19/England/QEUH-AA0484/2020\|EPI_ISL_665999\|20 | ···················· | ···················· | ···················· | ···················· | ···················· | ···················· |
| T | 5031 hCoV-19/England/CAMC-B78958/2020\|EPI_ISL_668101\|20 | ···················· | ···················· | ···················· | ···················· | ···················· | ···················· |
| T | 5032 hCoV-19/England/QEUH-B80F1F/2020\|EPI_ISL_668102\|20 | ···················· | ···················· | ···················· | ···················· | ···················· | ···················· |
| T | 5033 hCoV-19/England/QEUH-B7EA5F/2020\|EPI_ISL_668105\|20 | ···················· | ···················· | ···················· | ···················· | ···················· | ···················· |
| T | 5034 hCoV-19/England/QEUH-B7E5C7/2020\|EPI_ISL_668106\|20 | ···················· | ···················· | ···················· | ···················· | ···················· | ···················· |
| T | 5035 hCoV-19/England/QEUH-B80E6D/2020\|EPI_ISL_668107\|20 | ···················· | ···················· | ···················· | ···················· | ···················· | ···················· |
| T | 5036 hCoV-19/England/CAMC-B7A9CF/2020\|EPI_ISL_668110\|20 | ···················· | C··················· | ···················· | ···················· | ···················· | ···················· |
| T | 5037 hCoV-19/England/QEUH-B80E5E/2020\|EPI_ISL_668113\|20 | ···················· | ···················· | ···················· | ···················· | ···················· | ···················· |
| T | 5038 hCoV-19/England/QEUH-B7EE08/2020\|EPI_ISL_668114\|20 | ···················· | ···················· | ···················· | ···················· | ···················· | ···················· |
| T | 5039 hCoV-19/England/CAMC-B78BF8/2020\|EPI_ISL_668115\|20 | ···················· | ···················· | ···················· | ···················· | ···················· | ···················· |
| T | 5040 hCoV-19/England/QEUH-B7EE9F/2020\|EPI_ISL_668116\|20 | ···················· | ···················· | ···················· | ···················· | ···················· | ···················· |
| T | 5041 hCoV-19/England/QEUH-B81A68/2020\|EPI_ISL_668118\|20 | ···················· | ···················· | ···················· | ···················· | ···················· | ···················· |
| T | 5042 hCoV-19/England/QEUH-B7E916/2020\|EPI_ISL_668119\|20 | ···················· | ···················· | ···················· | ···················· | ···················· | ···················· |
| T | 5043 hCoV-19/Denmark/DCGC-10148/2020\|EPI_ISL_669418\|202 | ···················· | ···················· | ···················· | ···················· | ···················· | ···················· |
| T | 5044 hCoV-19/Denmark/DCGC-10149/2020\|EPI_ISL_669419\|202 | ···················· | ···················· | ···················· | ···················· | ···················· | ···················· |
| T | 5045 hCoV-19/Denmark/DCGC-10150/2020\|EPI_ISL_669420\|202 | ···················· | ···················· | ···················· | ···················· | ···················· | ···················· |
| T | 5046 hCoV-19/Denmark/DCGC-10151/2020\|EPI_ISL_669421\|202 | ···················· | ···················· | ···················· | ···················· | ···················· | ···················· |
| T | 5047 hCoV-19/Denmark/DCGC-10152/2020\|EPI_ISL_669422\|202 | ···················· | ···················· | ···················· | ···················· | ···················· | ···················· |
| T | 5048 hCoV-19/Denmark/DCGC-10153/2020\|EPI_ISL_669423\|202 | ···················· | ·A·················· | ···················· | ···················· | ···················· | ··A················· |

| | | | | | | | | |
|---|---|---|---|---|---|---|---|---|
| T | 5049 | hCoV-19/Denmark/DCGC-10154/2020\|EPI_ISL_669424\|202 | ···················· | ···················· | ····················· | ···················· | ···················· | ···················· |
| T | 5050 | hCoV-19/Denmark/DCGC-10164/2020\|EPI_ISL_669429\|202 | ···················· | ···················· | ····················· | ···················· | ···················· | ···················· |
| T | 5051 | hCoV-19/Denmark/DCGC-10165/2020\|EPI_ISL_669430\|202 | ···················· | ···················· | ····················· | ···················· | ···················· | ···················· |
| T | 5052 | hCoV-19/Denmark/DCGC-10167/2020\|EPI_ISL_669431\|202 | ···················· | ···················· | ····················· | ···················· | ···················· | ···················· |
| T | 5053 | hCoV-19/Denmark/DCGC-10168/2020\|EPI_ISL_669432\|202 | ···················· | ···················· | ····················· | ···················· | ···················· | ···················· |
| T | 5054 | hCoV-19/Denmark/DCGC-10169/2020\|EPI_ISL_669433\|202 | ···················· | ···················· | ····················· | ···················· | ···················· | ···················· |
| T | 5055 | hCoV-19/Denmark/DCGC-10170/2020\|EPI_ISL_669434\|202 | ···················· | ···················· | ····················· | ···················· | ···················· | ···················· |
| T | 5056 | hCoV-19/Denmark/DCGC-10172/2020\|EPI_ISL_669435\|202 | ···················· | ···················· | ····················· | ···················· | ···················· | ···················· |
| T | 5057 | hCoV-19/Denmark/DCGC-10173/2020\|EPI_ISL_669436\|202 | ···················· | ···················· | ····················· | ···················· | ···················· | ···················· |
| T | 5058 | hCoV-19/Denmark/DCGC-10174/2020\|EPI_ISL_669437\|202 | ···················· | ···················· | ····················· | ···················· | ···················· | ···················· |
| T | 5059 | hCoV-19/Denmark/DCGC-10175/2020\|EPI_ISL_669438\|202 | ···················· | ···················· | ····················· | ···················· | ···················· | ···················· |
| T | 5060 | hCoV-19/Denmark/DCGC-10176/2020\|EPI_ISL_669439\|202 | ···················· | ·A·················· | ····················· | ···················· | ···················· | ··A················· |
| T | 5061 | hCoV-19/Denmark/DCGC-10177/2020\|EPI_ISL_669440\|202 | ···················· | ···················· | ····················· | ···················· | ···················· | ···················· |
| T | 5062 | hCoV-19/Denmark/DCGC-10178/2020\|EPI_ISL_669441\|202 | ···················· | ···················· | ····················· | ···················· | ···················· | ···················· |
| T | 5063 | hCoV-19/Denmark/DCGC-10184/2020\|EPI_ISL_669442\|202 | ···················· | ·A·················· | ····················· | ···················· | ···················· | ··A················· |
| T | 5064 | hCoV-19/Denmark/DCGC-10190/2020\|EPI_ISL_669443\|202 | ···················· | ···················· | ····················· | ···················· | ···················· | ···················· |
| T | 5065 | hCoV-19/Denmark/DCGC-10191/2020\|EPI_ISL_669444\|202 | ···················· | ···················· | ····················· | ···················· | ···················· | ···················· |
| T | 5066 | hCoV-19/Denmark/DCGC-10192/2020\|EPI_ISL_669445\|202 | ···················· | ·A·················· | ····················· | ···················· | ···················· | ···················· |
| T | 5067 | hCoV-19/Denmark/DCGC-10194/2020\|EPI_ISL_669446\|202 | ···················· | ·A·················· | ····················· | ···················· | ···················· | ···················· |
| T | 5068 | hCoV-19/Denmark/DCGC-10196/2020\|EPI_ISL_669447\|202 | ···················· | ···················· | ····················· | ···················· | ···················· | ···················· |
| T | 5069 | hCoV-19/Denmark/DCGC-10197/2020\|EPI_ISL_669448\|202 | ···················· | ···················· | ····················· | ···················· | ···················· | ···················· |
| T | 5070 | hCoV-19/Denmark/DCGC-10198/2020\|EPI_ISL_669449\|202 | ···················· | ·A·················· | ····················· | ···················· | ···················· | ···················· |
| T | 5071 | hCoV-19/Denmark/DCGC-11117/2020\|EPI_ISL_670400\|202 | ···················· | ···················· | ····················· | ···················· | ···················· | ···················· |
| T | 5072 | hCoV-19/Denmark/DCGC-11119/2020\|EPI_ISL_670401\|202 | ···················· | ···················· | ····················· | ···················· | ···················· | ···················· |
| T | 5073 | hCoV-19/Denmark/DCGC-11121/2020\|EPI_ISL_670402\|202 | ···················· | ···················· | ····················· | ···················· | ···················· | ···················· |
| T | 5074 | hCoV-19/Denmark/DCGC-11122/2020\|EPI_ISL_670403\|202 | ···················· | ···················· | ····················· | ···················· | ······T············· | ···················· |
| T | 5075 | hCoV-19/Denmark/DCGC-11124/2020\|EPI_ISL_670404\|202 | ···················· | ···················· | ····················· | ···················· | ···················· | ···················· |
| T | 5076 | hCoV-19/Denmark/DCGC-11125/2020\|EPI_ISL_670405\|202 | ···················· | ···················· | ····················· | ···················· | ···················· | ···················· |
| T | 5077 | hCoV-19/Denmark/DCGC-11126/2020\|EPI_ISL_670406\|202 | ···················· | ···················· | ····················· | ···················· | ···················· | ···················· |
| T | 5078 | hCoV-19/Denmark/DCGC-11128/2020\|EPI_ISL_670407\|202 | ···················· | ···················· | ····················· | ···················· | ···················· | ···················· |
| T | 5079 | hCoV-19/Denmark/DCGC-11133/2020\|EPI_ISL_670408\|202 | ···················· | ···················· | ····················· | ···················· | ···················· | ···················· |
| T | 5080 | hCoV-19/Denmark/DCGC-11134/2020\|EPI_ISL_670409\|202 | ···················· | ···················· | ····················· | ···················· | ···················· | ···················· |
| T | 5081 | hCoV-19/Denmark/DCGC-11135/2020\|EPI_ISL_670410\|202 | ···················· | ···················· | ····················· | ···················· | ···················· | ···················· |
| T | 5082 | hCoV-19/Denmark/DCGC-11140/2020\|EPI_ISL_670411\|202 | ···················· | ···················· | ····················· | ···················· | ···················· | ···················· |
| T | 5083 | hCoV-19/Denmark/DCGC-11141/2020\|EPI_ISL_670412\|202 | ···················· | ···················· | ····················· | ···················· | ···················· | ···················· |
| T | 5084 | hCoV-19/Denmark/DCGC-11143/2020\|EPI_ISL_670413\|202 | ···················· | ···················· | ····················· | ···················· | ···················· | ···················· |
| T | 5085 | hCoV-19/Denmark/DCGC-11144/2020\|EPI_ISL_670414\|202 | ···················· | ·A·················· | ····················· | ···················· | ···················· | ···················· |
| T | 5086 | hCoV-19/Denmark/DCGC-11145/2020\|EPI_ISL_670415\|202 | ···················· | ···················· | ····················· | ···················· | ···················· | ···················· |
| T | 5087 | hCoV-19/Denmark/DCGC-11146/2020\|EPI_ISL_670416\|202 | ···················· | ···················· | ····················· | ···················· | ···················· | ···················· |
| T | 5088 | hCoV-19/Denmark/DCGC-11147/2020\|EPI_ISL_670417\|202 | ···················· | ···················· | ····················· | ···················· | ···················· | ···················· |
| T | 5089 | hCoV-19/Denmark/DCGC-11148/2020\|EPI_ISL_670418\|202 | ···················· | ···················· | ····················· | ···················· | ···················· | ···················· |
| T | 5090 | hCoV-19/Denmark/DCGC-11149/2020\|EPI_ISL_670419\|202 | ···················· | ···················· | ····················· | ···················· | ···················· | ···················· |
| T | 5091 | hCoV-19/Denmark/DCGC-11150/2020\|EPI_ISL_670420\|202 | ···················· | ···················· | ····················· | ···················· | ···················· | ···················· |
| T | 5092 | hCoV-19/Denmark/DCGC-11151/2020\|EPI_ISL_670421\|202 | ···················· | ···················· | ····················· | ···················· | ···················· | ···················· |
| T | 5093 | hCoV-19/Denmark/DCGC-11152/2020\|EPI_ISL_670422\|202 | ···················· | ···················· | ····················· | ···················· | ······T············· | ···················· |
| T | 5094 | hCoV-19/Denmark/DCGC-11154/2020\|EPI_ISL_670423\|202 | ···················· | ·A·················· | ····················· | ···················· | ···················· | ···················· |
| T | 5095 | hCoV-19/Denmark/DCGC-11155/2020\|EPI_ISL_670424\|202 | ···················· | ···················· | ····················· | ···················· | ···················· | ···················· |
| T | 5096 | hCoV-19/Denmark/DCGC-11157/2020\|EPI_ISL_670425\|202 | ···················· | ···················· | ····················· | ···················· | ···················· | ···················· |
| T | 5097 | hCoV-19/Denmark/DCGC-11158/2020\|EPI_ISL_670426\|202 | ···················· | ···················· | ····················· | ···················· | ···················· | ···················· |
| T | 5098 | hCoV-19/Denmark/DCGC-11159/2020\|EPI_ISL_670427\|202 | ···················· | ···················· | ····················· | ···················· | ···················· | ···················· |
| T | 5099 | hCoV-19/Denmark/DCGC-11163/2020\|EPI_ISL_670428\|202 | ···················· | ···················· | ····················· | ···················· | ···················· | ···················· |
| T | 5100 | hCoV-19/Denmark/DCGC-11165/2020\|EPI_ISL_670429\|202 | ···················· | ···················· | ····················· | ···················· | ···················· | ···················· |
| T | 5101 | hCoV-19/USA/NY-NYCPHL-001148/2020\|EPI_ISL_671730\|2 | ···················· | ···················· | ····················· | ···················· | ···················· | ···················· |
| T | 5102 | hCoV-19/USA/NY-NYCPHL-001150/2020\|EPI_ISL_671732\|2 | ···················· | ···················· | ····················· | ···················· | ···················· | ···················· |
| T | 5103 | hCoV-19/USA/NY-NYCPHL-001151/2020\|EPI_ISL_671733\|2 | ···················· | ·A·················· | ····················· | ···················· | ···················· | ···················· |
| T | 5104 | hCoV-19/USA/NY-NYCPHL-001153/2020\|EPI_ISL_671735\|2 | ···················· | ···················· | ····················· | ···················· | ···················· | ···················· |
| T | 5105 | hCoV-19/USA/NY-NYCPHL-001154/2020\|EPI_ISL_671736\|2 | ···················· | ···················· | ····················· | ···················· | ···················· | ···················· |
| T | 5106 | hCoV-19/USA/NY-NYCPHL-001157/2020\|EPI_ISL_671738\|2 | ···················· | ···················· | ····················· | ···················· | ···················· | ···················· |
| T | 5107 | hCoV-19/USA/NY-NYCPHL-001160/2020\|EPI_ISL_671740\|2 | ···················· | ···················· | ····················· | ···················· | ·······T············ | ···················· |
| T | 5108 | hCoV-19/USA/NY-NYCPHL-001164/2020\|EPI_ISL_671744\|2 | ···················· | ···················· | ····················· | ···················· | ···················· | ···················· |
| T | 5109 | hCoV-19/USA/NY-NYCPHL-001166/2020\|EPI_ISL_671746\|2 | ···················· | ···················· | ····················· | ···················· | ···················· | ···················· |
| T | 5110 | hCoV-19/USA/NY-NYCPHL-001167/2020\|EPI_ISL_671747\|2 | ···················· | ···················· | ····················· | ···················· | ···················· | ···················· |
| T | 5111 | hCoV-19/USA/NY-NYCPHL-001168/2020\|EPI_ISL_671748\|2 | ···················· | ···················· | ····················· | ···················· | ···················· | ···················· |
| T | 5112 | hCoV-19/USA/NY-NYCPHL-001169/2020\|EPI_ISL_671749\|2 | ···················· | ···················· | ····················· | ···················· | ···················· | ···················· |
| T | 5113 | hCoV-19/USA/NY-NYCPHL-001170/2020\|EPI_ISL_671750\|2 | ···················· | ···················· | ····················· | ···················· | ···················· | ···················· |
| T | 5114 | hCoV-19/USA/NY-NYCPHL-001171/2020\|EPI_ISL_671751\|2 | ···················· | ···················· | ····················· | ···················· | ···················· | ···················· |
| T | 5115 | hCoV-19/USA/NY-NYCPHL-001172/2020\|EPI_ISL_671752\|2 | ···················· | ···················· | ····················· | ···················· | ···················· | ···················· |
| T | 5116 | hCoV-19/USA/NY-NYCPHL-001173/2020\|EPI_ISL_671753\|2 | ···················· | ···················· | ····················· | ···················· | ···················· | ···················· |
| T | 5117 | hCoV-19/USA/NY-NYCPHL-001174/2020\|EPI_ISL_671754\|2 | ···················· | ···················· | ····················· | ···················· | ···················· | ···················· |
| T | 5118 | hCoV-19/USA/NY-NYCPHL-001178/2020\|EPI_ISL_671757\|2 | ···················· | ·A·················· | ····················· | ···················· | ···················· | ···················· |
| T | 5119 | hCoV-19/USA/NY-NYCPHL-001179/2020\|EPI_ISL_671758\|2 | ···················· | ···················· | ····················· | ···················· | ·······T············ | ···················· |
| T | 5120 | hCoV-19/USA/NY-NYCPHL-001181/2020\|EPI_ISL_671760\|2 | ···················· | ···················· | ····················· | ···················· | ···················· | ···················· |
| T | 5121 | hCoV-19/England/QEUH-B81B38/2020\|EPI_ISL_668010\|20 | ···················· | ···················· | ····················· | ···················· | ···················· | ···················· |
| T | 5122 | hCoV-19/England/CAMC-B78ABF/2020\|EPI_ISL_668015\|20 | ···················· | ···················· | ····················· | ···················· | ···················· | ···················· |
| T | 5123 | hCoV-19/England/CAMC-B791F5/2020\|EPI_ISL_668016\|20 | ···················· | ···················· | ····················· | ···················· | ······T············· | ···················· |
| T | 5124 | hCoV-19/England/QEUH-B7EF05/2020\|EPI_ISL_668021\|20 | ···················· | ···················· | ····················· | ···················· | ···················· | ···················· |
| T | 5125 | hCoV-19/England/QEUH-B80E9A/2020\|EPI_ISL_668022\|20 | ···················· | ···················· | ····················· | ···················· | ···················· | ···················· |
| T | 5126 | hCoV-19/England/CAMC-B79B42/2020\|EPI_ISL_668023\|20 | ···················· | ···················· | ····················· | ···················· | ···················· | ···················· |
| T | 5127 | hCoV-19/England/QEUH-B81AE0/2020\|EPI_ISL_668024\|20 | ···················· | ···················· | ····················· | ···················· | ···················· | ···················· |
| T | 5128 | hCoV-19/England/CAMC-B79210/2020\|EPI_ISL_668025\|20 | ···················· | ···················· | ····················· | ···················· | ···················· | ···················· |

T 5129 hCoV-19/England/CAMC-B79BAC/2020|EPI_ISL_668026|20 ···················· ·A·················· ···················· ···················· ···················· ···A················
T 5130 hCoV-19/England/CAMC-B78EE0/2020|EPI_ISL_668027|20 ···················· ···················· ···················· ···················· ···················· ····················
T 5131 hCoV-19/England/QEUH-B80F5B/2020|EPI_ISL_668028|20 ···················· ···················· ···················· ···················· ···················· ····················
T 5132 hCoV-19/England/QEUH-B81AC2/2020|EPI_ISL_668029|20 ···················· ···················· ···················· ···················· ···················· ····················
T 5133 hCoV-19/England/QEUH-B7EA40/2020|EPI_ISL_668030|20 ···················· ···················· ···················· ···················· ···················· ····················
T 5134 hCoV-19/England/QEUH-B7EECC/2020|EPI_ISL_668031|20 ···················· ···················· ···················· ···················· ···················· ····················
T 5135 hCoV-19/England/QEUH-B819F2/2020|EPI_ISL_668033|20 ···················· ···················· ···················· ···················· ···················· ····················
T 5136 hCoV-19/England/QEUH-B7E49D/2020|EPI_ISL_668034|20 ···················· ···················· ···················· ···················· ···················· ····················
T 5137 hCoV-19/England/QEUH-B819D4/2020|EPI_ISL_668035|20 ···················· ···················· ···················· ···················· ···················· ····················
T 5138 hCoV-19/England/CAMC-B78D98/2020|EPI_ISL_668037|20 ···················· ···················· ···················· ···················· ···················· ····················
T 5139 hCoV-19/England/CAMC-B789FE/2020|EPI_ISL_668041|20 ···················· ···················· ···················· ···················· ···················· ····················
T 5140 hCoV-19/England/CAMC-B799FD/2020|EPI_ISL_668042|20 ···················· ···················· ···················· ···················· ···················· ····················
T 5141 hCoV-19/England/CAMC-B78E2C/2020|EPI_ISL_668043|20 ···················· ···················· ···················· ···················· ···················· ····················
T 5142 hCoV-19/England/QEUH-B819A7/2020|EPI_ISL_668044|20 ···················· ···················· ···················· ···················· ···················· ····················
T 5143 hCoV-19/England/QEUH-B7E9CB/2020|EPI_ISL_668045|20 ···················· ···················· ···················· ···················· ···················· ····················
T 5144 hCoV-19/England/CAMC-B78D89/2020|EPI_ISL_668046|20 ···················· ···················· ···················· ···················· ···················· ····················
T 5145 hCoV-19/England/CAMC-B7B07E/2020|EPI_ISL_668048|20 ···················· ···················· ···················· ···················· ···················· ····················
T 5146 hCoV-19/England/QEUH-B8197A/2020|EPI_ISL_668049|20 ···················· ···················· ···················· ···················· ···················· ····················
T 5147 hCoV-19/England/QEUH-B7E952/2020|EPI_ISL_668050|20 ···················· ···················· ···················· ···················· ···················· ····················
T 5148 hCoV-19/England/QEUH-B7E970/2020|EPI_ISL_668052|20 ···················· ···················· ···················· ···················· ···················· ····················
T 5149 hCoV-19/England/QEUH-B7E503/2020|EPI_ISL_668053|20 ···················· ···················· ···················· ···················· ···················· ····················
T 5150 hCoV-19/England/QEUH-B7E925/2020|EPI_ISL_668054|20 ···················· ···················· ···················· ···················· ···················· ····················
T 5151 hCoV-19/England/QEUH-B7EB89/2020|EPI_ISL_668055|20 ···················· ···················· ···················· ···················· ···················· ····················
T 5152 hCoV-19/England/QEUH-B7EB98/2020|EPI_ISL_668056|20 ···················· ···················· ···················· ···················· ···················· ····················
T 5153 hCoV-19/England/QEUH-B7F12C/2020|EPI_ISL_668057|20 ···················· ···················· ···················· ···················· ···················· ····················
T 5154 hCoV-19/England/CAMC-B7916E/2020|EPI_ISL_668058|20 ···················· ···················· ···················· ···················· ···················· ····················
T 5155 hCoV-19/England/CAMC-B79B51/2020|EPI_ISL_668059|20 ···················· ···················· ···················· ···················· ···················· ····················
T 5156 hCoV-19/England/QEUH-B7E4CA/2020|EPI_ISL_668060|20 ···················· ···················· ···················· ···················· ···················· ····················
T 5157 hCoV-19/England/CAMC-B78F65/2020|EPI_ISL_668061|20 ···················· ···················· ···················· ···················· ···················· ····················
T 5158 hCoV-19/England/QEUH-B80F2E/2020|EPI_ISL_668062|20 ···················· ···················· ···················· ···················· ···················· ····················
T 5159 hCoV-19/England/QEUH-B8178F/2020|EPI_ISL_668063|20 ···················· ···················· ···················· ···················· ···················· ····················
T 5160 hCoV-19/England/QEUH-B7E4E8/2020|EPI_ISL_668064|20 ···················· ···················· ···················· ···················· ···················· ····················
T 5161 hCoV-19/England/CAMC-B79D4C/2020|EPI_ISL_668065|20 ···················· ···················· ···················· ···················· ···················· ····················
T 5162 hCoV-19/England/QEUH-B7E9F8/2020|EPI_ISL_668066|20 ···················· ···················· ···················· ···················· ···················· ····················
T 5163 hCoV-19/England/QEUH-B81A3B/2020|EPI_ISL_668067|20 ···················· ···················· ···················· ···················· ···················· ····················
T 5164 hCoV-19/England/CAMC-B78ED1/2020|EPI_ISL_668068|20 ···················· ···················· ···················· ···················· ···················· ····················
T 5165 hCoV-19/England/CAMC-B790CB/2020|EPI_ISL_668069|20 ···················· ···················· ···················· ···················· ···················· ····················
T 5166 hCoV-19/England/CAMC-B7A1D6/2020|EPI_ISL_668071|20 ···················· ···················· ···················· ···················· ···················· ····················
T 5167 hCoV-19/England/CAMC-B79A54/2020|EPI_ISL_668072|20 ···················· ···················· ···················· ···················· ···················· ····················
T 5168 hCoV-19/England/QEUH-B80F97/2020|EPI_ISL_668073|20 ···················· ···················· ···················· ···················· ···················· ····················
T 5169 hCoV-19/England/QEUH-B7EF23/2020|EPI_ISL_668074|20 ···················· ···················· ···················· ···················· ···················· ····················
T 5170 hCoV-19/England/QEUH-B7EB5C/2020|EPI_ISL_668075|20 ···················· ···················· ···················· ···················· ···················· ····················
T 5171 hCoV-19/England/CAMC-B7B0BA/2020|EPI_ISL_668076|20 ···················· ···················· ···················· ···················· ···················· ····················
T 5172 hCoV-19/England/CAMC-B78A37/2020|EPI_ISL_668077|20 ···················· ···················· ···················· ···················· ···················· ····················
T 5173 hCoV-19/England/QEUH-B81B0B/2020|EPI_ISL_668078|20 ···················· ···················· ···················· ···················· ···················· ····················
T 5174 hCoV-19/Denmark/DCGC-10020/2020|EPI_ISL_669340|202 ···················· ···················· ···················· ···················· ···················· ····················
T 5175 hCoV-19/Denmark/DCGC-10021/2020|EPI_ISL_669341|202 ···················· ···················· ···················· ···················· ···················· ····················
T 5176 hCoV-19/Denmark/DCGC-10022/2020|EPI_ISL_669342|202 ···················· ···················· ···················· ···················· ···················· ····················
T 5177 hCoV-19/Denmark/DCGC-10025/2020|EPI_ISL_669343|202 ···················· ···················· ···················· ···················· ···················· ····················
T 5178 hCoV-19/Denmark/DCGC-10028/2020|EPI_ISL_669346|202 ···················· ···················· ···················· ···················· ···················· ····················
T 5179 hCoV-19/Denmark/DCGC-10029/2020|EPI_ISL_669347|202 ···················· ···················· ···················· ···················· ······T············· ····················
T 5180 hCoV-19/Denmark/DCGC-10035/2020|EPI_ISL_669350|202 ···················· ···················· ···················· ···················· ···················· ····················
T 5181 hCoV-19/Denmark/DCGC-10036/2020|EPI_ISL_669351|202 ···················· ···················· ···················· ···················· ···················· ····················
T 5182 hCoV-19/Denmark/DCGC-10037/2020|EPI_ISL_669352|202 ···················· ···················· ···················· ···················· ···················· ····················
T 5183 hCoV-19/Denmark/DCGC-10040/2020|EPI_ISL_669353|202 ···················· ····A··············· ···················· ···················· ···················· ····················
T 5184 hCoV-19/Denmark/DCGC-10041/2020|EPI_ISL_669354|202 ···················· ···················· ···················· ···················· ···················· ····················
T 5185 hCoV-19/Denmark/DCGC-10042/2020|EPI_ISL_669355|202 ···················· ···················· ···················· ···················· ···················· ····················
T 5186 hCoV-19/Denmark/DCGC-10043/2020|EPI_ISL_669356|202 ···················· ·A·················· ···················· ···················· ···················· ···A················
T 5187 hCoV-19/Denmark/DCGC-10044/2020|EPI_ISL_669357|202 ···················· ···················· ···················· ···················· ···················· ····················
T 5188 hCoV-19/Denmark/DCGC-10045/2020|EPI_ISL_669358|202 ···················· ···················· ···················· ···················· ···················· ····················
T 5189 hCoV-19/Denmark/DCGC-10046/2020|EPI_ISL_669359|202 ···················· ···················· ···················· ···················· ···················· ····················
T 5190 hCoV-19/Denmark/DCGC-10048/2020|EPI_ISL_669360|202 ···················· ···················· ···················· ···················· ···················· ····················
T 5191 hCoV-19/Denmark/DCGC-10049/2020|EPI_ISL_669361|202 ···················· ···················· ···················· ···················· ···················· ····················
T 5192 hCoV-19/Denmark/DCGC-10051/2020|EPI_ISL_669362|202 ···················· ···················· ···················· ···················· ···················· ····················
T 5193 hCoV-19/Denmark/DCGC-10053/2020|EPI_ISL_669363|202 ···················· ···················· ···················· ···················· ···················· ····················
T 5194 hCoV-19/Denmark/DCGC-10054/2020|EPI_ISL_669364|202 ···················· ···················· ···················· ···················· ···················· ····················
T 5195 hCoV-19/Denmark/DCGC-10055/2020|EPI_ISL_669365|202 ···················· ···················· ···················· ···················· ···················· ····················
T 5196 hCoV-19/Denmark/DCGC-10056/2020|EPI_ISL_669366|202 ···················· ···················· ···················· ···················· ···················· ····················
T 5197 hCoV-19/Denmark/DCGC-10057/2020|EPI_ISL_669367|202 ···················· ·A·················· ···················· ···················· ···················· ···A················
T 5198 hCoV-19/Denmark/DCGC-10058/2020|EPI_ISL_669368|202 ···················· ···················· ···················· ···················· ···················· ····················
T 5199 hCoV-19/Denmark/DCGC-10059/2020|EPI_ISL_669369|202 ···················· ···················· ···················· ···················· ···················· ····················
T 5200 hCoV-19/Denmark/DCGC-10061/2020|EPI_ISL_669370|202 ···················· ···················· ···················· ···················· ······T············· ····················
T 5201 hCoV-19/Denmark/DCGC-10062/2020|EPI_ISL_669371|202 ···················· ···················· ···················· ···················· ···················· ····················
T 5202 hCoV-19/Denmark/DCGC-10063/2020|EPI_ISL_669372|202 ···················· ···················· ···················· ···················· ···················· ····················
T 5203 hCoV-19/Denmark/DCGC-10064/2020|EPI_ISL_669373|202 ···················· ···················· ···················· ···················· ···················· ····················
T 5204 hCoV-19/Denmark/DCGC-10066/2020|EPI_ISL_669374|202 ···················· ···················· ···················· ···················· ···················· ····················
T 5205 hCoV-19/Denmark/DCGC-10067/2020|EPI_ISL_669375|202 ···················· ···················· ···················· ···················· ···················· ····················
T 5206 hCoV-19/Denmark/DCGC-10068/2020|EPI_ISL_669376|202 ···················· ···················· ···················· ···················· ···················· ····················
T 5207 hCoV-19/Denmark/DCGC-10071/2020|EPI_ISL_669377|202 ···················· ···················· ···················· ···················· ···················· ····················
T 5208 hCoV-19/Denmark/DCGC-10072/2020|EPI_ISL_669378|202 ···················· ···················· ···················· ···················· ···················· ····················

T 5209 hCoV-19/Denmark/DCGC-10074/2020|EPI_ISL_669379|202 ···················· ···················· ···················· ···················· ···················· ····················
T 5210 hCoV-19/Denmark/DCGC-10075/2020|EPI_ISL_669380|202 ···················· ···················· ···················· ···················· ·····T·············· ····················
T 5211 hCoV-19/Denmark/DCGC-10077/2020|EPI_ISL_669381|202 ···················· ·A·················· ···················· ···················· ···················· ··A·················
T 5212 hCoV-19/Denmark/DCGC-10080/2020|EPI_ISL_669382|202 ···················· ···················· ···················· ···················· ···················· ····················
T 5213 hCoV-19/Denmark/DCGC-10081/2020|EPI_ISL_669383|202 ···················· ···················· ···················· ···················· ···················· ····················
T 5214 hCoV-19/Denmark/DCGC-10083/2020|EPI_ISL_669384|202 ···················· ···················· ···················· ···················· ···················· ····················
T 5215 hCoV-19/Denmark/DCGC-10084/2020|EPI_ISL_669385|202 ···················· ···················· ···················· ···················· ···················· ····················
T 5216 hCoV-19/Denmark/DCGC-10088/2020|EPI_ISL_669386|202 ···················· ···················· ···················· ···················· ···················· ····················
T 5217 hCoV-19/Denmark/DCGC-10089/2020|EPI_ISL_669387|202 ···················· ···A················ ···················· ···················· ···················· ····················
T 5218 hCoV-19/Denmark/DCGC-10092/2020|EPI_ISL_669388|202 ···················· ···················· ···················· ···················· ···················· ····················
T 5219 hCoV-19/Denmark/DCGC-10093/2020|EPI_ISL_669389|202 ···················· ···················· ···················· ···················· ···················· ····················
T 5220 hCoV-19/Denmark/DCGC-10094/2020|EPI_ISL_669390|202 ···················· ···················· ···················· ···················· ···················· ····················
T 5221 hCoV-19/Denmark/DCGC-10097/2020|EPI_ISL_669391|202 ···················· ···················· ···················· ···················· ···················· ····················
T 5222 hCoV-19/Denmark/DCGC-10098/2020|EPI_ISL_669392|202 ···················· ···················· ···················· ···················· ···················· ····················
T 5223 hCoV-19/Denmark/DCGC-10100/2020|EPI_ISL_669393|202 ···················· ···················· ···················· ···················· ···················· ····················
T 5224 hCoV-19/Denmark/DCGC-10102/2020|EPI_ISL_669394|202 ···················· ·A·················· ···················· ···················· ···················· ····················
T 5225 hCoV-19/Denmark/DCGC-10104/2020|EPI_ISL_669395|202 ···················· ···················· ···················· ···················· ···················· ····················
T 5226 hCoV-19/Denmark/DCGC-10105/2020|EPI_ISL_669396|202 ···················· ·A·················· ···················· ···················· ···················· ··A·················
T 5227 hCoV-19/Denmark/DCGC-10106/2020|EPI_ISL_669397|202 ···················· ···················· ···················· ···················· ···················· ····················
T 5228 hCoV-19/Denmark/DCGC-10108/2020|EPI_ISL_669398|202 ···················· ···················· ···················· ···················· ···················· ····················
T 5229 hCoV-19/Denmark/DCGC-10109/2020|EPI_ISL_669399|202 ···················· ·A·················· ···················· ···················· ···················· ····················
T 5230 hCoV-19/Denmark/DCGC-10111/2020|EPI_ISL_669400|202 ···················· ···················· ···················· ···················· ···················· ····················
T 5231 hCoV-19/Denmark/DCGC-10112/2020|EPI_ISL_669401|202 ···················· ···················· ···················· ···················· ···················· ····················
T 5232 hCoV-19/Denmark/DCGC-10113/2020|EPI_ISL_669402|202 ···················· ···················· ···················· ···················· ···················· ····················
T 5233 hCoV-19/Denmark/DCGC-10114/2020|EPI_ISL_669403|202 ···················· ···················· ···················· ···················· ···················· ····················
T 5234 hCoV-19/Denmark/DCGC-10115/2020|EPI_ISL_669404|202 ···················· ···················· ···················· ···················· ···················· ····················
T 5235 hCoV-19/Denmark/DCGC-10116/2020|EPI_ISL_669405|202 ···················· ···················· ···················· ···················· ···················· ····················
T 5236 hCoV-19/Denmark/DCGC-10117/2020|EPI_ISL_669406|202 ···················· ···················· ···················· ···················· ···················· ····················
T 5237 hCoV-19/Denmark/DCGC-10120/2020|EPI_ISL_669407|202 ···················· ···················· ···················· ···················· ···················· ····················
T 5238 hCoV-19/Denmark/DCGC-10121/2020|EPI_ISL_669408|202 ···················· ···················· ···················· ···················· ···················· ····················
T 5239 hCoV-19/Denmark/DCGC-10123/2020|EPI_ISL_669409|202 ···················· ·A·················· ···················· ···················· ···················· ····················
T 5240 hCoV-19/Denmark/DCGC-10128/2020|EPI_ISL_669410|202 ···················· ···················· ···················· ···················· ···················· ····················
T 5241 hCoV-19/Denmark/DCGC-10130/2020|EPI_ISL_669411|202 ···················· ···················· ···················· ···················· ···················· ····················
T 5242 hCoV-19/Denmark/DCGC-10138/2020|EPI_ISL_669412|202 ···················· ···················· ···················· ···················· ···················· ····················
T 5243 hCoV-19/Denmark/DCGC-10141/2020|EPI_ISL_669413|202 ···················· ···················· ···················· ···················· ···················· ····················
T 5244 hCoV-19/Denmark/DCGC-10142/2020|EPI_ISL_669414|202 ···················· ···················· ···················· ···················· ···················· ····················
T 5245 hCoV-19/Denmark/DCGC-10143/2020|EPI_ISL_669415|202 ···················· ···················· ···················· ···················· ···················· ····················
T 5246 hCoV-19/Denmark/DCGC-10144/2020|EPI_ISL_669416|202 ···················· ···················· ···················· ···················· ···················· ····················
T 5247 hCoV-19/Denmark/DCGC-10147/2020|EPI_ISL_669417|202 ···················· ···················· ···················· ···················· ···················· ····················
T 5248 hCoV-19/Denmark/DCGC-10155/2020|EPI_ISL_669425|202 ···················· ···················· ···················· ···················· ···················· ····················
T 5249 hCoV-19/Denmark/DCGC-10156/2020|EPI_ISL_669426|202 ···················· ···················· ···················· ···················· ···················· ····················
T 5250 hCoV-19/Denmark/DCGC-10160/2020|EPI_ISL_669427|202 ···················· ···················· ···················· ···················· ···················· ····················
T 5251 hCoV-19/Denmark/DCGC-10163/2020|EPI_ISL_669428|202 ···················· ·A·················· ···················· ···················· ···················· ····················
T 5252 hCoV-19/Denmark/DCGC-10976/2020|EPI_ISL_670320|202 ···················· ···················· ···················· ···················· ···················· ····················
T 5253 hCoV-19/Denmark/DCGC-10977/2020|EPI_ISL_670321|202 ···················· ···················· ···················· ···················· ···················· ····················
T 5254 hCoV-19/Denmark/DCGC-10979/2020|EPI_ISL_670322|202 ···················· ···················· ···················· ···················· ···················· ····················
T 5255 hCoV-19/Denmark/DCGC-10980/2020|EPI_ISL_670323|202 ···················· ···················· ···················· ···················· ···················· ····················
T 5256 hCoV-19/Denmark/DCGC-10981/2020|EPI_ISL_670324|202 ···················· ···················· ···················· ···················· ···················· ····················
T 5257 hCoV-19/Denmark/DCGC-10982/2020|EPI_ISL_670325|202 ···················· ·A·················· ···················· ···················· ···················· ··A·················
T 5258 hCoV-19/Denmark/DCGC-10983/2020|EPI_ISL_670326|202 ···················· ···················· ···················· ···················· ···················· ····················
T 5259 hCoV-19/Denmark/DCGC-10985/2020|EPI_ISL_670327|202 ···················· ···················· ···················· ···················· ···················· ····················
T 5260 hCoV-19/Denmark/DCGC-10986/2020|EPI_ISL_670328|202 ···················· ···················· ···················· ···················· ···················· ····················
T 5261 hCoV-19/Denmark/DCGC-10988/2020|EPI_ISL_670329|202 ···················· ···················· ···················· ···················· ···················· ····················
T 5262 hCoV-19/Denmark/DCGC-10991/2020|EPI_ISL_670330|202 ···················· ···················· ···················· ···················· ···················· ····················
T 5263 hCoV-19/Denmark/DCGC-10994/2020|EPI_ISL_670331|202 ···················· ·A·················· ···················· ···················· ···················· ····················
T 5264 hCoV-19/Denmark/DCGC-10995/2020|EPI_ISL_670332|202 ···················· ···················· ···················· ···················· ···················· ····················
T 5265 hCoV-19/Denmark/DCGC-10996/2020|EPI_ISL_670333|202 ···················· ···················· ···················· ···················· ···················· ····················
T 5266 hCoV-19/Denmark/DCGC-10998/2020|EPI_ISL_670334|202 ···················· ···················· ···················· ···················· ···················· ····················
T 5267 hCoV-19/Denmark/DCGC-11000/2020|EPI_ISL_670335|202 ···················· ···················· ···················· ···················· ···················· ····················
T 5268 hCoV-19/Denmark/DCGC-11001/2020|EPI_ISL_670336|202 ···················· ···················· ···················· ···················· ···················· ····················
T 5269 hCoV-19/Denmark/DCGC-11002/2020|EPI_ISL_670337|202 ···················· ···················· ···················· ···················· ···················· ····················
T 5270 hCoV-19/Denmark/DCGC-11003/2020|EPI_ISL_670338|202 ···················· ···················· ···················· ···················· ···················· ····················
T 5271 hCoV-19/Denmark/DCGC-11006/2020|EPI_ISL_670339|202 ···················· ···················· ···················· ···················· ···················· ····················
T 5272 hCoV-19/Denmark/DCGC-11007/2020|EPI_ISL_670340|202 ···················· ···················· ···················· ···················· ···················· ····················
T 5273 hCoV-19/Denmark/DCGC-11008/2020|EPI_ISL_670341|202 ···················· ·A·················· ···················· ···················· ···················· ··A·················
T 5274 hCoV-19/Denmark/DCGC-11010/2020|EPI_ISL_670342|202 ···················· ···················· ···················· ···················· ···················· ····················
T 5275 hCoV-19/Denmark/DCGC-11011/2020|EPI_ISL_670343|202 ···················· ···················· ···················· ···················· ···················· ····················
T 5276 hCoV-19/Denmark/DCGC-11012/2020|EPI_ISL_670344|202 ···················· ···················· ···················· ···················· ···················· ····················
T 5277 hCoV-19/Denmark/DCGC-11015/2020|EPI_ISL_670345|202 ···················· ···················· ···················· ···················· ···················· ····················
T 5278 hCoV-19/Denmark/DCGC-11017/2020|EPI_ISL_670346|202 ···················· ···················· ···················· ···················· ···················· ····················
T 5279 hCoV-19/Denmark/DCGC-11018/2020|EPI_ISL_670347|202 ···················· ···················· ···················· ···················· ···················· ····················
T 5280 hCoV-19/Denmark/DCGC-11019/2020|EPI_ISL_670348|202 ···················· ···················· ···················· ···················· ···················· ····················
T 5281 hCoV-19/Denmark/DCGC-11023/2020|EPI_ISL_670349|202 ···················· ···················· ···················· ···················· ···················· ····················
T 5282 hCoV-19/Denmark/DCGC-11024/2020|EPI_ISL_670350|202 ···················· ···················· ···················· ···················· ···················· ····················
T 5283 hCoV-19/Denmark/DCGC-11027/2020|EPI_ISL_670351|202 ···················· ···················· ···················· ···················· ···················· ····················
T 5284 hCoV-19/Denmark/DCGC-11028/2020|EPI_ISL_670352|202 ···················· ···················· ···················· ···················· ···················· ····················
T 5285 hCoV-19/Denmark/DCGC-11029/2020|EPI_ISL_670353|202 ···················· ···················· ···················· ···················· ···················· ····················
T 5286 hCoV-19/Denmark/DCGC-11030/2020|EPI_ISL_670354|202 ···················· ···················· ···················· ···················· ···················· ····················
T 5287 hCoV-19/Denmark/DCGC-11031/2020|EPI_ISL_670355|202 ···················· ···················· ···················· ···················· ···················· ····················
T 5288 hCoV-19/Denmark/DCGC-11032/2020|EPI_ISL_670356|202 ···················· ···················· ···················· ···················· ···················· ····················

T 5289 hCoV-19/Denmark/DCGC-11036/2020|EPI_ISL_670357|202 ···················· ···················· ···················· ···················· ···················· ····················
T 5290 hCoV-19/Denmark/DCGC-11037/2020|EPI_ISL_670358|202 ···················· ···················· ···················· ···················· ···················· ····················
T 5291 hCoV-19/Denmark/DCGC-11039/2020|EPI_ISL_670359|202 ···················· ···················· ···················· ···················· ···················· ····················
T 5292 hCoV-19/Denmark/DCGC-11040/2020|EPI_ISL_670360|202 ···················· ···················· ···················· ···················· ···················· ····················
T 5293 hCoV-19/Denmark/DCGC-11042/2020|EPI_ISL_670361|202 ···················· ···················· ···················· ···················· ···················· ····················
T 5294 hCoV-19/Denmark/DCGC-11043/2020|EPI_ISL_670362|202 ···················· ···················· ···················· ···················· ···················· ····················
T 5295 hCoV-19/Denmark/DCGC-11045/2020|EPI_ISL_670363|202 ···················· ···················· ···················· ···················· ···················· ····················
T 5296 hCoV-19/Denmark/DCGC-11047/2020|EPI_ISL_670364|202 ···················· ···················· ···················· ···················· ···················· ····················
T 5297 hCoV-19/Denmark/DCGC-11050/2020|EPI_ISL_670365|202 ···················· ···················· ···················· ···················· ···················· ····················
T 5298 hCoV-19/Denmark/DCGC-11051/2020|EPI_ISL_670366|202 ···················· ···················· ···················· ···················· ···················· ····················
T 5299 hCoV-19/Denmark/DCGC-11052/2020|EPI_ISL_670367|202 ···················· ···················· ···················· ···················· ···················· ····················
T 5300 hCoV-19/Denmark/DCGC-11054/2020|EPI_ISL_670368|202 ···················· ···················· ···················· ···················· ···················· ····················
T 5301 hCoV-19/Denmark/DCGC-11057/2020|EPI_ISL_670369|202 ···················· ·A·················· ···················· ···················· ···················· ····················
T 5302 hCoV-19/Denmark/DCGC-11059/2020|EPI_ISL_670370|202 ···················· ···················· ···················· ···················· ···················· ····················
T 5303 hCoV-19/Denmark/DCGC-11060/2020|EPI_ISL_670371|202 ···················· ···················· ···················· ···················· ···················· ····················
T 5304 hCoV-19/Denmark/DCGC-11062/2020|EPI_ISL_670372|202 ···················· ···················· ···················· ···················· ···················· ····················
T 5305 hCoV-19/Denmark/DCGC-11063/2020|EPI_ISL_670373|202 ···················· ···················· ···················· ···················· ···················· ····················
T 5306 hCoV-19/Denmark/DCGC-11064/2020|EPI_ISL_670374|202 ···················· ···················· ···················· ···················· ···················· ····················
T 5307 hCoV-19/Denmark/DCGC-11065/2020|EPI_ISL_670375|202 ···················· ···················· ···················· ···················· ···················· ····················
T 5308 hCoV-19/Denmark/DCGC-11067/2020|EPI_ISL_670376|202 ···················· ···················· ···················· ···················· ···················· ····················
T 5309 hCoV-19/Denmark/DCGC-11069/2020|EPI_ISL_670377|202 ···················· ···················· ···················· ···················· ···················· ····················
T 5310 hCoV-19/Denmark/DCGC-11070/2020|EPI_ISL_670378|202 ···················· ···················· ···················· ···················· ···················· ····················
T 5311 hCoV-19/Denmark/DCGC-11074/2020|EPI_ISL_670379|202 ···················· ···················· ···················· ···················· ···················· ····················
T 5312 hCoV-19/Denmark/DCGC-11077/2020|EPI_ISL_670380|202 ···················· ···················· ···················· ···················· ···················· ····················
T 5313 hCoV-19/Denmark/DCGC-11078/2020|EPI_ISL_670381|202 ···················· ···················· ···················· ···················· ···················· ····················
T 5314 hCoV-19/Denmark/DCGC-11081/2020|EPI_ISL_670382|202 ···················· ···················· ···················· ···················· ···················· ····················
T 5315 hCoV-19/Denmark/DCGC-11087/2020|EPI_ISL_670383|202 ···················· ···················· ···················· ···················· ···················· ····················
T 5316 hCoV-19/Denmark/DCGC-11088/2020|EPI_ISL_670384|202 ···················· ···················· ···················· ···················· ···················· ····················
T 5317 hCoV-19/Denmark/DCGC-11089/2020|EPI_ISL_670385|202 ···················· ·A·················· ···················· ···················· ···················· ····················
T 5318 hCoV-19/Denmark/DCGC-11090/2020|EPI_ISL_670386|202 ···················· ···················· ···················· ···················· ···················· ····················
T 5319 hCoV-19/Denmark/DCGC-11092/2020|EPI_ISL_670387|202 ···················· ···················· ···················· ···················· ···················· ····················
T 5320 hCoV-19/Denmark/DCGC-11093/2020|EPI_ISL_670388|202 ···················· ·A·················· ···················· ···················· ···················· ····················
T 5321 hCoV-19/Denmark/DCGC-11094/2020|EPI_ISL_670389|202 ···················· ···················· ···················· ···················· ···················· ····················
T 5322 hCoV-19/Denmark/DCGC-11096/2020|EPI_ISL_670390|202 ···················· ···················· ···················· ···················· ···················· ····················
T 5323 hCoV-19/Denmark/DCGC-11097/2020|EPI_ISL_670391|202 ···················· ···················· ···················· ···················· ···················· ····················
T 5324 hCoV-19/Denmark/DCGC-11101/2020|EPI_ISL_670392|202 ···················· ···················· ···················· ···················· ···················· ····················
T 5325 hCoV-19/Denmark/DCGC-11103/2020|EPI_ISL_670393|202 ···················· ······C············· ···················· ···················· ···················· ····················
T 5326 hCoV-19/Denmark/DCGC-11105/2020|EPI_ISL_670394|202 ···················· ···················· ···················· ···················· ···················· ····················
T 5327 hCoV-19/Denmark/DCGC-11106/2020|EPI_ISL_670395|202 ···················· ···················· ···················· ···················· ···················· ····················
T 5328 hCoV-19/Denmark/DCGC-11108/2020|EPI_ISL_670396|202 ···················· ···················· ···················· ···················· ···················· ····················
T 5329 hCoV-19/Canada/ON-UHTC-0362/2020|EPI_ISL_671652|20 ···················· ···················· ···················· ···················· ···················· ····················
T 5330 hCoV-19/Canada/ON-UHTC-0316/2020|EPI_ISL_671653|20 ···················· ···················· ···················· ···················· ···················· ····················
T 5331 hCoV-19/Canada/ON-UHTC-0375/2020|EPI_ISL_671654|20 ···················· ·A·················· ···················· ···················· ···················· ··A·················
T 5332 hCoV-19/Canada/ON-UHTC-0363/2020|EPI_ISL_671655|20 ···················· ···················· ···················· ···················· ···················· ····················
T 5333 hCoV-19/Canada/ON-UHTC-0366/2020|EPI_ISL_671656|20 ···················· ···················· ···················· ···················· ···················· ····················
T 5334 hCoV-19/Canada/ON-UHTC-0372/2020|EPI_ISL_671657|20 ···················· ·A·················· ···················· ···················· ···················· ··A·················
T 5335 hCoV-19/Canada/ON-UHTC-0374/2020|EPI_ISL_671658|20 ···················· ···················· ···················· ···················· ···················· ····················
T 5336 hCoV-19/Canada/ON-UHTC-0376/2020|EPI_ISL_671659|20 ···················· ···················· ···················· ···················· ···················· ····················
T 5337 hCoV-19/Canada/ON-UHTC-0369/2020|EPI_ISL_671660|20 ···················· ···················· ···················· ···················· ···················· ····················
T 5338 hCoV-19/Canada/ON-UHTC-0360/2020|EPI_ISL_671663|20 ···················· ···················· ···················· ···················· ···················· ····················
T 5339 hCoV-19/Canada/ON-UHTC-0373/2020|EPI_ISL_671665|20 ···················· ···················· ···················· ···················· ···················· ····················
T 5340 hCoV-19/Canada/ON-UHTC-0365/2020|EPI_ISL_671666|20 ···················· ·A·················· ···················· ···················· ···················· ··A·················
T 5341 hCoV-19/Canada/ON-UHTC-0377/2020|EPI_ISL_671667|20 ···················· ···················· ···················· ···················· ···················· ····················
T 5342 hCoV-19/Canada/ON-UHTC-0364/2020|EPI_ISL_671668|20 ···················· ···················· ···················· ···················· ···················· ····················
T 5343 hCoV-19/Canada/ON-UHTC-0367/2020|EPI_ISL_671669|20 ···················· ·A·················· ···················· ···················· ···················· ··A·················
T 5344 hCoV-19/Canada/ON-UHTC-0361/2020|EPI_ISL_671670|20 ···················· ···················· ···················· ···················· ···················· ····················
T 5345 hCoV-19/Canada/ON-UHTC-0368/2020|EPI_ISL_671671|20 ···················· ···················· ···················· ···················· ···················· ····················
T 5346 hCoV-19/Australia/NSW1191/2020|EPI_ISL_671673|2020 ···················· ·A·················· ···················· ···················· ···················· ··A·················
T 5347 hCoV-19/USA/NY-NYCPHL-001091/2020|EPI_ISL_671682|2 ···················· ···················· ···················· ···················· ···················· ····················
T 5348 hCoV-19/USA/NY-NYCPHL-001093/2020|EPI_ISL_671683|2 ···················· ···················· ···················· ···················· ···················· ····················
T 5349 hCoV-19/USA/NY-NYCPHL-001094/2020|EPI_ISL_671684|2 ···················· ···················· ···················· ···················· ···················· ····················
T 5350 hCoV-19/USA/NY-NYCPHL-001100/2020|EPI_ISL_671685|2 ···················· ···················· ···················· ···················· ···················· ····················
T 5351 hCoV-19/USA/NY-NYCPHL-001104/2020|EPI_ISL_671687|2 ···················· ···················· ···················· ···················· ···················· ····················
T 5352 hCoV-19/USA/NY-NYCPHL-001107/2020|EPI_ISL_671690|2 ···················· ···················· ···················· ···················· ···················· ····················
T 5353 hCoV-19/USA/NY-NYCPHL-001109/2020|EPI_ISL_671692|2 ···················· ···················· ···················· ···················· ···················· ····················
T 5354 hCoV-19/USA/NY-NYCPHL-001110/2020|EPI_ISL_671693|2 ···················· ···················· ···················· ···················· ···················· ····················
T 5355 hCoV-19/USA/NY-NYCPHL-001112/2020|EPI_ISL_671695|2 ···················· ···················· ···················· ···················· ···················· ····················
T 5356 hCoV-19/USA/NY-NYCPHL-001115/2020|EPI_ISL_671698|2 ···················· ·A·················· ···················· ···················· ···················· ····················
T 5357 hCoV-19/USA/NY-NYCPHL-001116/2020|EPI_ISL_671699|2 ···················· ···················· ···················· ···················· ···················· ····················
T 5358 hCoV-19/USA/NY-NYCPHL-001117/2020|EPI_ISL_671700|2 ···················· ···················· ···················· ···················· ···················· ····················
T 5359 hCoV-19/USA/NY-NYCPHL-001118/2020|EPI_ISL_671701|2 ···················· ···················· ···················· ···················· ···················· ····················
T 5360 hCoV-19/USA/NY-NYCPHL-001120/2020|EPI_ISL_671703|2 ···················· ···················· ···················· ···················· ···················· ····················
T 5361 hCoV-19/USA/NY-NYCPHL-001126/2020|EPI_ISL_671709|2 ···················· ···················· ···················· ···················· ···················· ····················
T 5362 hCoV-19/USA/NY-NYCPHL-001127/2020|EPI_ISL_671710|2 ···················· ···················· ···················· ···················· ···················· ····················
T 5363 hCoV-19/USA/NY-NYCPHL-001129/2020|EPI_ISL_671712|2 ···················· ···················· ···················· ···················· ···················· ····················
T 5364 hCoV-19/USA/NY-NYCPHL-001130/2020|EPI_ISL_671713|2 ···················· ···················· ···················· ···················· ···················· ····················
T 5365 hCoV-19/USA/NY-NYCPHL-001132/2020|EPI_ISL_671715|2 ···················· ···················· ···················· ···················· ···················· ····················
T 5366 hCoV-19/USA/NY-NYCPHL-001133/2020|EPI_ISL_671716|2 ···················· ···················· ···················· ···················· ···················· ····················
T 5367 hCoV-19/USA/NY-NYCPHL-001136/2020|EPI_ISL_671719|2 ···················· ···················· ···················· ···················· ···················· ····················
T 5368 hCoV-19/USA/NY-NYCPHL-001137/2020|EPI_ISL_671720|2 ···················· ···················· ···················· ···················· ···················· ····················

T 5369 hCoV-19/USA/NY-NYCPHL-001138/2020|EPI_ISL_671721|2 ···················· ···················· ···················· ···················· ···················· ····················
T 5370 hCoV-19/USA/NY-NYCPHL-001139/2020|EPI_ISL_671722|2 ···················· ···················· ···················· ···················· ···················· ····················
T 5371 hCoV-19/USA/NY-NYCPHL-001140/2020|EPI_ISL_671723|2 ···················· ·A·················· ···················· ···················· ···················· ····················
T 5372 hCoV-19/USA/NY-NYCPHL-001141/2020|EPI_ISL_671724|2 ···················· ···················· ···················· ···················· ···················· ····················
T 5373 hCoV-19/USA/NY-NYCPHL-001142/2020|EPI_ISL_671725|2 ···················· ···················· ···················· ···················· ···················· ····················
T 5374 hCoV-19/USA/NY-NYCPHL-001144/2020|EPI_ISL_671726|2 ···················· ···················· ···················· ···················· ···················· ····················
T 5375 hCoV-19/USA/NY-NYCPHL-001145/2020|EPI_ISL_671727|2 ···················· ·A·················· ···················· ···················· ···················· ····················
T 5376 hCoV-19/USA/NY-NYCPHL-001146/2020|EPI_ISL_671728|2 ···················· ···················· ···················· ···················· ···················· ····················
T 5377 hCoV-19/England/QEUH-AF5505/2020|EPI_ISL_663104|20 ···················· ···················· ···················· ···················· ···················· ····················
T 5378 hCoV-19/England/QEUH-AF4BC8/2020|EPI_ISL_663106|20 ···················· ···················· ···················· ···················· ···················· ····················
T 5379 hCoV-19/England/QEUH-AF52C2/2020|EPI_ISL_663109|20 ···················· ···················· ···················· ···················· ···················· ····················
T 5380 hCoV-19/England/QEUH-AF613D/2020|EPI_ISL_663110|20 ···················· ···················· ···················· ···················· ···················· ····················
T 5381 hCoV-19/England/QEUH-AF5ABB/2020|EPI_ISL_663111|20 ···················· ···················· ···················· ···················· ···················· ····················
T 5382 hCoV-19/England/QEUH-AF5365/2020|EPI_ISL_663112|20 ···················· ···················· ···················· ···················· ···················· ····················
T 5383 hCoV-19/England/QEUH-AF5620/2020|EPI_ISL_663115|20 ···················· ·A·················· ···················· ···················· ···················· ···A················
T 5384 hCoV-19/England/QEUH-AF40AB/2020|EPI_ISL_663116|20 ···················· ···················· ···················· ···················· ···················· ····················
T 5385 hCoV-19/England/QEUH-AF49CE/2020|EPI_ISL_663117|20 ···················· ···················· ···················· ···················· ····T··············· ····················
T 5386 hCoV-19/England/QEUH-AF6971/2020|EPI_ISL_663118|20 ···················· ···················· ···················· ···················· ···················· ····················
T 5387 hCoV-19/England/QEUH-AF5532/2020|EPI_ISL_663119|20 ···················· ···················· ···················· ···················· ···················· ····················
T 5388 hCoV-19/England/QEUH-AF517A/2020|EPI_ISL_663120|20 ···················· ···················· ···················· ···················· ···················· ····················
T 5389 hCoV-19/England/QEUH-AF4946/2020|EPI_ISL_663121|20 ···················· ···················· ···················· ···················· ···················· ····················
T 5390 hCoV-19/England/QEUH-AF57A5/2020|EPI_ISL_663123|20 ···················· ···················· ···················· ···················· ···················· ····················
T 5391 hCoV-19/England/QEUH-AF5B03/2020|EPI_ISL_663124|20 ···················· ···················· ···················· ···················· ···················· ····················
T 5392 hCoV-19/England/QEUH-AF4C3E/2020|EPI_ISL_663126|20 ···················· ···················· ···················· ···················· ···················· ····················
T 5393 hCoV-19/England/QEUH-AF582A/2020|EPI_ISL_663127|20 ···················· ·A·················· ···················· ···················· ···················· ····················
T 5394 hCoV-19/England/QEUH-AF573C/2020|EPI_ISL_663129|20 ···················· ···················· ···················· ···················· ···················· ····················
T 5395 hCoV-19/England/QEUH-AF4C5C/2020|EPI_ISL_663130|20 ···················· ···················· ···················· ···················· ···················· ····················
T 5396 hCoV-19/England/QEUH-AF5B30/2020|EPI_ISL_663131|20 ···················· ···················· ···················· ···················· ···················· ····················
T 5397 hCoV-19/England/QEUH-AF5408/2020|EPI_ISL_663132|20 ···················· ···················· ···················· ···················· ···················· ····················
T 5398 hCoV-19/England/QEUH-AF48EF/2020|EPI_ISL_663133|20 ···················· ···················· ···················· ···················· ···················· ····················
T 5399 hCoV-19/England/QEUH-AF5338/2020|EPI_ISL_663134|20 ···················· ···················· ···················· ···················· ···················· ····················
T 5400 hCoV-19/England/QEUH-AF4B9B/2020|EPI_ISL_663135|20 ···················· ···················· ···················· ···················· ···················· ····················
T 5401 hCoV-19/England/QEUH-AF5ACA/2020|EPI_ISL_663136|20 ···················· ···················· ···················· ···················· ···················· ····················
T 5402 hCoV-19/England/QEUH-AF4E0B/2020|EPI_ISL_663137|20 ···················· ···················· ···················· ···················· ···················· ····················
T 5403 hCoV-19/England/QEUH-AF4CF2/2020|EPI_ISL_663138|20 ···················· ···················· ···················· ···················· ···················· ····················
T 5404 hCoV-19/England/QEUH-AF5866/2020|EPI_ISL_663139|20 ···················· ···················· ···················· ···················· ···················· ····················
T 5405 hCoV-19/England/QEUH-AF51C5/2020|EPI_ISL_663140|20 ···················· ···················· ···················· ···················· ···················· ····················
T 5406 hCoV-19/England/QEUH-AF4982/2020|EPI_ISL_663141|20 ···················· ···················· ···················· ···················· ···················· ····················
T 5407 hCoV-19/England/QEUH-AF6A23/2020|EPI_ISL_663142|20 ···················· ···················· ···················· ···················· ···················· ····················
T 5408 hCoV-19/England/QEUH-AF566C/2020|EPI_ISL_663143|20 ···················· ···················· ···················· ···················· ···················· ····················
T 5409 hCoV-19/England/QEUH-AF490A/2020|EPI_ISL_663144|20 ···················· ·A·················· ···················· ···················· ···················· ···A················
T 5410 hCoV-19/England/QEUH-AF5374/2020|EPI_ISL_663145|20 ···················· ···················· ···················· ···················· ······T············· ····················
T 5411 hCoV-19/England/QEUH-AF4B40/2020|EPI_ISL_663146|20 ···················· ···················· ···················· ···················· ···················· ····················
T 5412 hCoV-19/England/QEUH-AF4991/2020|EPI_ISL_663147|20 ···················· ·A·················· ···················· ···················· ···················· ···A················
T 5413 hCoV-19/England/QEUH-AF51D4/2020|EPI_ISL_663148|20 ···················· ···················· ···················· ···················· ···················· ····················
T 5414 hCoV-19/England/QEUH-AF4BD7/2020|EPI_ISL_663149|20 ···················· ···················· ···················· ···················· ···················· ····················
T 5415 hCoV-19/England/QEUH-AF3D78/2020|EPI_ISL_663150|20 ···················· ·A·················· ···················· ···················· ···················· ····················
T 5416 hCoV-19/England/QEUH-AF3D0F/2020|EPI_ISL_663151|20 ···················· ···················· ···················· ···················· ···················· ····················
T 5417 hCoV-19/England/QEUH-AF66D4/2020|EPI_ISL_663152|20 ···················· ···················· ···················· ···················· ···················· ····················
T 5418 hCoV-19/England/QEUH-AF4B31/2020|EPI_ISL_663153|20 ···················· ···················· ···················· ···················· ···················· ····················
T 5419 hCoV-19/England/QEUH-AF48C1/2020|EPI_ISL_663154|20 ···················· ···················· ···················· ···················· ···················· ····················
T 5420 hCoV-19/England/QEUH-AF69EA/2020|EPI_ISL_663155|20 ···················· ···················· ···················· ···················· ···················· ····················
T 5421 hCoV-19/England/QEUH-AF5295/2020|EPI_ISL_663156|20 ···················· ·A·················· ···················· ···················· ···················· ···A················
T 5422 hCoV-19/England/QEUH-AF6917/2020|EPI_ISL_663157|20 ···················· ···················· ···················· ···················· ···················· ····················
T 5423 hCoV-19/England/QEUH-AF4E47/2020|EPI_ISL_663158|20 ···················· ···················· ···················· ···················· ···················· ····················
T 5424 hCoV-19/England/QEUH-AF5DD0/2020|EPI_ISL_663160|20 ···················· ···················· ···················· ···················· ···················· ····················
T 5425 hCoV-19/England/QEUH-AF6944/2020|EPI_ISL_663161|20 ···················· ·A·················· ···················· ···················· ···················· ····················
T 5426 hCoV-19/England/QEUH-AF55C9/2020|EPI_ISL_663162|20 ···················· ·A·················· ···················· ···················· ···················· ····················
T 5427 hCoV-19/England/QEUH-AF575A/2020|EPI_ISL_663163|20 ···················· ···················· ···················· ···················· ···················· ····················
T 5428 hCoV-19/England/QEUH-AF5462/2020|EPI_ISL_663164|20 ···················· ···················· ···················· ···················· ···················· ····················
T 5429 hCoV-19/England/QEUH-AF521D/2020|EPI_ISL_663165|20 ···················· ···················· ···················· ···················· ···················· ····················
T 5430 hCoV-19/England/QEUH-AF4AE9/2020|EPI_ISL_663166|20 ···················· ···················· ···················· ···················· ···················· ····················
T 5431 hCoV-19/England/QEUH-AF6908/2020|EPI_ISL_663167|20 ···················· ···················· ···················· ···················· ···················· ····················
T 5432 hCoV-19/England/QEUH-AF5198/2020|EPI_ISL_663168|20 ···················· ···················· ···················· ···················· ···················· ····················
T 5433 hCoV-19/England/QEUH-AF5453/2020|EPI_ISL_663169|20 ···················· ···················· ···················· ···················· ···················· ····················
T 5434 hCoV-19/England/QEUH-AF5EDD/2020|EPI_ISL_663170|20 ···················· ···················· ···················· ···················· ···················· ····················
T 5435 hCoV-19/England/QEUH-AF5BF4/2020|EPI_ISL_663172|20 ···················· ···················· ···················· ···················· ···················· ···A················
T 5436 hCoV-19/England/QEUH-AF563F/2020|EPI_ISL_663173|20 ···················· ···················· ···················· ···················· ···················· ····················
T 5437 hCoV-19/England/QEUH-AF413F/2020|EPI_ISL_663174|20 ···················· ···················· ···················· ···················· ···················· ····················
T 5438 hCoV-19/England/QEUH-AF530B/2020|EPI_ISL_663175|20 ···················· ···················· ···················· ···················· ···················· ····················
T 5439 hCoV-19/England/QEUH-AF4A07/2020|EPI_ISL_663176|20 ···················· ···················· ···················· ···················· ···················· ····················
T 5440 hCoV-19/England/QEUH-AF4A52/2020|EPI_ISL_663177|20 ···················· ···················· ···················· ···················· ···················· ····················
T 5441 hCoV-19/England/QEUH-AF5426/2020|EPI_ISL_663180|20 ···················· ···················· ···················· ···················· ···················· ····················
T 5442 hCoV-19/England/QEUH-AF53DE/2020|EPI_ISL_663181|20 ···················· ···················· ···················· ···················· ···················· ····················
T 5443 hCoV-19/England/QEUH-AF54EA/2020|EPI_ISL_663182|20 ···················· ···················· ···················· ···················· ···················· ····················
T 5444 hCoV-19/England/QEUH-AF3E39/2020|EPI_ISL_663183|20 ···················· ···················· ···················· ···················· ···················· ····················
T 5445 hCoV-19/England/QEUH-AF5B7C/2020|EPI_ISL_663184|20 ···················· ···················· ···················· ···················· ···················· ···A················
T 5446 hCoV-19/England/QEUH-AF66E3/2020|EPI_ISL_663186|20 ···················· ···················· ···················· ···················· ···················· ····················
T 5447 hCoV-19/England/QEUH-AF581B/2020|EPI_ISL_663187|20 ···················· ·A·················· ···················· ···················· ···················· ····················
T 5448 hCoV-19/England/QEUH-AF3E2A/2020|EPI_ISL_663188|20 ···················· ···················· ···················· ···················· ···················· ····················

| | | | | | | | |
|---|---|---|---|---|---|---|---|
| T | 5449 hCoV-19/England/QEUH-AF61D3/2020\|EPI_ISL_663189\|20 | ···················· | ···················· | ····················· | ···················· | ···················· | ···················· |
| T | 5450 hCoV-19/England/QEUH-AF55D8/2020\|EPI_ISL_663201\|20 | ···················· | ···················· | ····················· | ···················· | ···················· | ···················· |
| T | 5451 hCoV-19/England/QEUH-AF4B5F/2020\|EPI_ISL_663202\|20 | ···················· | ···················· | ····················· | ···················· | ···················· | ···················· |
| T | 5452 hCoV-19/England/QEUH-AF3E0C/2020\|EPI_ISL_663203\|20 | ···················· | ·A·················· | ····················· | ···················· | ···················· | ···················· |
| T | 5453 hCoV-19/England/QEUH-AF53B0/2020\|EPI_ISL_663204\|20 | ···················· | ···················· | ····················· | ···················· | ···················· | ···················· |
| T | 5454 hCoV-19/England/QEUH-AF5B12/2020\|EPI_ISL_663205\|20 | ···················· | ···················· | ····················· | ···················· | ···················· | ···················· |
| T | 5455 hCoV-19/France/NAQ-SC252/2020\|EPI_ISL_663206\|2020- | ···················· | ···················· | ····················· | ···················· | ···················· | ···················· |
| T | 5456 hCoV-19/France/NAQ-SC253/2020\|EPI_ISL_663207\|2020- | ···················· | ···················· | ····················· | ···················· | ···················· | ···················· |
| T | 5457 hCoV-19/France/NAQ-SC257/2020\|EPI_ISL_663208\|2020- | ···················· | ···················· | ····················· | ···················· | ···················· | ···················· |
| T | 5458 hCoV-19/France/NAQ-SC259/2020\|EPI_ISL_663209\|2020- | ···················· | ···················· | ····················· | ···················· | ···················· | ···················· |
| T | 5459 hCoV-19/France/NAQ-SC260/2020\|EPI_ISL_663210\|2020- | ···················· | ···················· | ····················· | ···················· | ···················· | ···················· |
| T | 5460 hCoV-19/France/NAQ-SC261/2020\|EPI_ISL_663211\|2020- | ···················· | ···················· | ····················· | ···················· | ···················· | ···················· |
| T | 5461 hCoV-19/France/NAQ-SC262/2020\|EPI_ISL_663212\|2020- | ···················· | ···················· | ····················· | ···················· | ···················· | ···················· |
| T | 5462 hCoV-19/France/NAQ-SC263/2020\|EPI_ISL_663213\|2020- | ···················· | ···················· | ····················· | ···················· | ···················· | ···················· |
| T | 5463 hCoV-19/France/NAQ-SC264/2020\|EPI_ISL_663214\|2020- | ···················· | ···················· | ····················· | ···················· | ···················· | ···················· |
| T | 5464 hCoV-19/France/NAQ-SC265/2020\|EPI_ISL_663215\|2020- | ···················· | ···················· | ····················· | ···················· | ···················· | ···················· |
| T | 5465 hCoV-19/France/NAQ-SC267/2020\|EPI_ISL_663216\|2020- | ···················· | ···················· | ····················· | ···················· | ···················· | ···················· |
| T | 5466 hCoV-19/France/NAQ-SC268/2020\|EPI_ISL_663217\|2020- | ···················· | ···················· | ····················· | ···················· | ···················· | ···················· |
| T | 5467 hCoV-19/France/NAQ-SC269/2020\|EPI_ISL_663218\|2020- | ···················· | ···················· | ····················· | ···················· | ···················· | ···················· |
| T | 5468 hCoV-19/France/NAQ-SC270/2020\|EPI_ISL_663219\|2020- | ···················· | ···················· | ····················· | ···················· | ···················· | ···················· |
| T | 5469 hCoV-19/France/NAQ-SC271/2020\|EPI_ISL_663220\|2020- | ···················· | ···················· | ····················· | ···················· | ···················· | ···················· |
| T | 5470 hCoV-19/France/NAQ-SC272/2020\|EPI_ISL_663221\|2020- | ···················· | ···················· | ····················· | ···················· | ···················· | ···················· |
| T | 5471 hCoV-19/France/NAQ-SC275/2020\|EPI_ISL_663222\|2020- | ···················· | ···················· | ····················· | ···················· | ···················· | ···················· |
| T | 5472 hCoV-19/France/NAQ-SC276/2020\|EPI_ISL_663223\|2020- | ···················· | ···················· | ····················· | ···················· | ···················· | ···················· |
| T | 5473 hCoV-19/France/NAQ-SC285/2020\|EPI_ISL_663224\|2020- | ···················· | ···················· | ····················· | ···················· | ···················· | ···················· |
| T | 5474 hCoV-19/Wales/PHWC-48B203/2020\|EPI_ISL_665766\|2020 | ···················· | ···················· | ····················· | ···················· | ···················· | ···················· |
| T | 5475 hCoV-19/Wales/PHWC-48B485/2020\|EPI_ISL_665767\|2020 | ···················· | ···················· | ····················· | ···················· | ···················· | ···················· |
| T | 5476 hCoV-19/Wales/PHWC-48B5A0/2020\|EPI_ISL_665768\|2020 | ···················· | ···················· | ····················· | ···················· | ···················· | ···················· |
| T | 5477 hCoV-19/Wales/PHWC-48BA38/2020\|EPI_ISL_665769\|2020 | ···················· | ···················· | ····················· | ···················· | ···················· | ···················· |
| T | 5478 hCoV-19/Wales/PHWC-48BE3C/2020\|EPI_ISL_665770\|2020 | ···················· | ···················· | ····················· | ···················· | ···················· | ···················· |
| T | 5479 hCoV-19/Wales/PHWC-48BE69/2020\|EPI_ISL_665771\|2020 | ···················· | ···················· | ····················· | ···················· | ···················· | ···················· |
| T | 5480 hCoV-19/Wales/PHWC-48BE87/2020\|EPI_ISL_665772\|2020 | ···················· | ···················· | ····················· | ···················· | ···················· | ···················· |
| T | 5481 hCoV-19/Wales/PHWC-48C387/2020\|EPI_ISL_665775\|2020 | ···················· | ···················· | ····················· | ···················· | ···················· | ···················· |
| T | 5482 hCoV-19/Wales/PHWC-48C484/2020\|EPI_ISL_665776\|2020 | ···················· | ···················· | ····················· | ···················· | ···················· | ···················· |
| T | 5483 hCoV-19/Wales/PHWC-48C5AF/2020\|EPI_ISL_665777\|2020 | ···················· | ···················· | ····················· | ···················· | ···················· | ···················· |
| T | 5484 hCoV-19/Wales/PHWC-48CACE/2020\|EPI_ISL_665778\|2020 | ···················· | ···················· | ····················· | ···················· | ···················· | ···················· |
| T | 5486 hCoV-19/Wales/PHWC-48CEB3/2020\|EPI_ISL_665780\|2020 | ···················· | ···················· | ····················· | ···················· | ···················· | ···················· |
| T | 5487 hCoV-19/Wales/PHWC-48CED1/2020\|EPI_ISL_665781\|2020 | ···················· | ···················· | ····················· | ···················· | ···················· | ···················· |
| T | 5488 hCoV-19/England/LOND-1250EB7/2020\|EPI_ISL_665785\|2 | ···················· | ···················· | ····················· | ···················· | ···················· | ···················· |
| T | 5489 hCoV-19/England/LOND-1251153/2020\|EPI_ISL_665786\|2 | ···················· | ···················· | ····················· | ···················· | ···················· | ···················· |
| T | 5490 hCoV-19/England/NORT-28D063/2020\|EPI_ISL_665787\|20 | ···················· | ···················· | ····················· | ···················· | ···················· | ···················· |
| T | 5492 hCoV-19/England/NOTT-11CEBB/2020\|EPI_ISL_665789\|20 | ···················· | ·A·················· | ····················· | ···················· | ···················· | ··A················· |
| T | 5493 hCoV-19/Scotland/EDB9320/2020\|EPI_ISL_665790\|2020- | ···················· | ···················· | ····················· | ···················· | ···················· | ···················· |
| T | 5494 hCoV-19/Scotland/EDB9321/2020\|EPI_ISL_665791\|2020- | ···················· | ···················· | ····················· | ···················· | ···················· | ···················· |
| T | 5496 hCoV-19/England/LOND-1250D14/2020\|EPI_ISL_665801\|2 | ···················· | ···················· | ····················· | ···················· | ···················· | ···················· |
| T | 5497 hCoV-19/Wales/PHWC-48C17E/2020\|EPI_ISL_665802\|2020 | ···················· | ···················· | ····················· | ···················· | ···················· | ···················· |
| T | 5498 hCoV-19/Wales/PHWC-48C994/2020\|EPI_ISL_665803\|2020 | ···················· | ···················· | ····················· | ···················· | ···················· | ···················· |
| T | 5499 hCoV-19/England/LOND-1250BDE/2020\|EPI_ISL_665805\|2 | ···················· | ···················· | ····················· | ···················· | ···················· | ···················· |
| T | 5500 hCoV-19/England/LOND-1250BFC/2020\|EPI_ISL_665806\|2 | ···················· | ···················· | ····················· | ···················· | ·····T·············· | ···················· |
| T | 5502 hCoV-19/England/LOND-1250CDB/2020\|EPI_ISL_665808\|2 | ···················· | ···················· | ····················· | ···················· | ·····T·············· | ···················· |
| T | 5503 hCoV-19/England/LOND-1250CF9/2020\|EPI_ISL_665809\|2 | ···················· | ···················· | ····················· | ···················· | ·····T·············· | ···················· |
| T | 5504 hCoV-19/England/LOND-1250D05/2020\|EPI_ISL_665810\|2 | ···················· | ···················· | ····················· | ···················· | ·····T·············· | ···················· |
| T | 5505 hCoV-19/England/LOND-1250D32/2020\|EPI_ISL_665811\|2 | ···················· | ···················· | ····················· | ···················· | ···················· | ···················· |
| T | 5506 hCoV-19/England/LOND-1250D41/2020\|EPI_ISL_665812\|2 | ···················· | ···················· | ····················· | ···················· | ···················· | ···················· |
| T | 5507 hCoV-19/England/LOND-1250D7E/2020\|EPI_ISL_665813\|2 | ···················· | ···················· | ····················· | ···················· | ·····T·············· | ···················· |
| T | 5508 hCoV-19/England/LOND-1250DAB/2020\|EPI_ISL_665814\|2 | ···················· | ···················· | ····················· | ···················· | ···················· | ···················· |
| T | 5509 hCoV-19/England/LOND-1250DC9/2020\|EPI_ISL_665815\|2 | ···················· | ···················· | ····················· | ···················· | ···················· | ···················· |
| T | 5510 hCoV-19/England/LOND-1250DE7/2020\|EPI_ISL_665816\|2 | ···················· | ···················· | ····················· | ···················· | ···················· | ···················· |
| T | 5513 hCoV-19/England/LOND-1250E20/2020\|EPI_ISL_665819\|2 | ···················· | ···················· | ····················· | ···················· | ···················· | ···················· |
| T | 5514 hCoV-19/England/LOND-1250E3F/2020\|EPI_ISL_665820\|2 | ···················· | ···················· | ····················· | ···················· | ···················· | ···················· |
| T | 5516 hCoV-19/England/LOND-1250E6C/2020\|EPI_ISL_665822\|2 | ···················· | ···················· | ····················· | ···················· | ···················· | ···················· |
| T | 5517 hCoV-19/England/LOND-1250F2D/2020\|EPI_ISL_665823\|2 | ···················· | ···················· | ····················· | ···················· | ···················· | ···················· |
| T | 5518 hCoV-19/England/LOND-1250F78/2020\|EPI_ISL_665824\|2 | ···················· | ···················· | ····················· | ···················· | ···················· | ···················· |
| T | 5519 hCoV-19/England/LOND-1250F96/2020\|EPI_ISL_665825\|2 | ···················· | ···················· | ····················· | ···················· | ···················· | ···················· |
| T | 5521 hCoV-19/England/LOND-125101A/2020\|EPI_ISL_665827\|2 | ···················· | ···················· | ····················· | ···················· | ···················· | ···················· |
| T | 5523 hCoV-19/England/LOND-1251056/2020\|EPI_ISL_665829\|2 | ···················· | ···················· | ····················· | ···················· | ·····T·············· | ···················· |
| T | 5524 hCoV-19/England/LOND-1251065/2020\|EPI_ISL_665830\|2 | ···················· | ·A·················· | ····················· | ···················· | ···················· | ··A················· |
| T | 5525 hCoV-19/England/LOND-1251108/2020\|EPI_ISL_665831\|2 | ···················· | ···················· | ····················· | ···················· | ···················· | ···················· |
| T | 5526 hCoV-19/England/LOND-1251171/2020\|EPI_ISL_665832\|2 | ···················· | ···················· | ····················· | ···················· | ···················· | ···················· |
| T | 5527 hCoV-19/England/LOND-1251180/2020\|EPI_ISL_665833\|2 | ···················· | ···················· | ····················· | ···················· | ···················· | ···················· |
| T | 5528 hCoV-19/England/LOND-12511BD/2020\|EPI_ISL_665834\|2 | ···················· | ···················· | ····················· | ···················· | ·····T·············· | ···················· |
| T | 5529 hCoV-19/England/LOND-12511CC/2020\|EPI_ISL_665835\|2 | ···················· | ···················· | ····················· | ···················· | ···················· | ···················· |
| T | 5531 hCoV-19/England/LOND-1251205/2020\|EPI_ISL_665837\|2 | ···················· | ···················· | ····················· | ···················· | ·····T·············· | ···················· |
| T | 5532 hCoV-19/England/LOND-1251214/2020\|EPI_ISL_665838\|2 | ···················· | ···················· | ····················· | ···················· | ···················· | ···················· |
| T | 5534 hCoV-19/England/LOND-1251250/2020\|EPI_ISL_665840\|2 | ···················· | ···················· | ····················· | ···················· | ···················· | ···················· |
| T | 5535 hCoV-19/England/LOND-125126F/2020\|EPI_ISL_665841\|2 | ···················· | ···················· | ····················· | ···················· | ···················· | ···················· |
| T | 5536 hCoV-19/England/LOND-12512BA/2020\|EPI_ISL_665842\|2 | ···················· | ···················· | ····················· | ···················· | ···················· | ···················· |
| T | 5537 hCoV-19/England/LOND-12512F6/2020\|EPI_ISL_665843\|2 | ···················· | ···················· | ····················· | ···················· | ···················· | ···················· |
| T | 5539 hCoV-19/England/LOND-125136C/2020\|EPI_ISL_665845\|2 | ···················· | ···················· | ····················· | ···················· | ···················· | ···················· |
| T | 5540 hCoV-19/England/LOND-12513A8/2020\|EPI_ISL_665846\|2 | ···················· | ···················· | ····················· | ···················· | ···················· | ···················· |

| | | | | | | | |
|---|---|---|---|---|---|---|---|
| T | 5541 hCoV-19/England/LOND-12513E4/2020\|EPI_ISL_665847\|2 | ···················· | ···················· | ····················· | ···················· | ···················· | ···················· |
| T | 5542 hCoV-19/England/LOND-125140F/2020\|EPI_ISL_665848\|2 | ···················· | ···················· | ····················· | ···················· | ···················· | ···················· |
| T | 5543 hCoV-19/England/MILK-A23CEB/2020\|EPI_ISL_665849\|20 | ···················· | ···················· | ····················· | ···················· | ···················· | ···················· |
| T | 5546 hCoV-19/England/MILK-A697CA/2020\|EPI_ISL_665852\|20 | ···················· | ···················· | ····················· | ···················· | ···················· | ···················· |
| T | 5548 hCoV-19/England/MILK-A6988B/2020\|EPI_ISL_665854\|20 | ···················· | ···················· | ····················· | ···················· | ···················· | ···················· |
| T | 5552 hCoV-19/England/MILK-A69A58/2020\|EPI_ISL_665858\|20 | ···················· | ···················· | ····················· | ···················· | ···················· | ···················· |
| T | 5554 hCoV-19/England/MILK-A69A85/2020\|EPI_ISL_665860\|20 | ···················· | ···················· | ····················· | ···················· | ···················· | ···················· |
| T | 5555 hCoV-19/England/MILK-A69AC1/2020\|EPI_ISL_665861\|20 | ···················· | ···················· | ····················· | ···················· | ···················· | ···················· |
| T | 5556 hCoV-19/England/MILK-A69B19/2020\|EPI_ISL_665862\|20 | ···················· | ···················· | ····················· | ···················· | ···················· | ···················· |
| T | 5558 hCoV-19/England/MILK-B05194/2020\|EPI_ISL_665864\|20 | ···················· | ···················· | ····················· | ···················· | ···················· | ···················· |
| T | 5559 hCoV-19/England/MILK-B065F1/2020\|EPI_ISL_665865\|20 | ···················· | ···················· | ····················· | ···················· | ···················· | ···················· |
| T | 5560 hCoV-19/England/MILK-B06D08/2020\|EPI_ISL_665866\|20 | ···················· | ···················· | ····················· | ···················· | ···················· | ···················· |
| T | 5561 hCoV-19/England/MILK-B06E7E/2020\|EPI_ISL_665867\|20 | ···················· | ···················· | ····················· | ···················· | ···················· | ···················· |
| T | 5562 hCoV-19/England/NORT-28CD12/2020\|EPI_ISL_665868\|20 | ···················· | ···················· | ····················· | ···················· | ···················· | ···················· |
| T | 5563 hCoV-19/England/NORT-28CD6D/2020\|EPI_ISL_665869\|20 | ···················· | ···················· | ····················· | ···················· | ···················· | ···················· |
| T | 5564 hCoV-19/England/NORT-28CD8B/2020\|EPI_ISL_665870\|20 | ···················· | ···················· | ····················· | ···················· | ···················· | ···················· |
| T | 5565 hCoV-19/England/NORT-28CD9A/2020\|EPI_ISL_665871\|20 | ···················· | ···················· | ····················· | ···················· | ···················· | ···················· |
| T | 5566 hCoV-19/England/NORT-28CDD6/2020\|EPI_ISL_665872\|20 | ···················· | ···················· | ····················· | ···················· | ···················· | ···················· |
| T | 5567 hCoV-19/England/NORT-28D054/2020\|EPI_ISL_665873\|20 | ···················· | ···················· | ····················· | ···················· | ···················· | ···················· |
| T | 5568 hCoV-19/England/NORT-28D072/2020\|EPI_ISL_665874\|20 | ···················· | ···················· | ····················· | ···················· | ···················· | ···················· |
| T | 5569 hCoV-19/England/NORT-28D081/2020\|EPI_ISL_665875\|20 | ···················· | ···················· | ····················· | ···················· | ···················· | ···················· |
| T | 5570 hCoV-19/England/NORW-F2CC1/2020\|EPI_ISL_665876\|202 | ···················· | ···················· | ····················· | ···················· | ···················· | ···················· |
| T | 5571 hCoV-19/England/NORW-F4E9C/2020\|EPI_ISL_665877\|202 | ···················· | ···················· | ····················· | ···················· | ······T············· | ···················· |
| T | 5573 hCoV-19/England/NORW-F4F99/2020\|EPI_ISL_665879\|202 | ···················· | ···················· | ····················· | ···················· | ···················· | ···················· |
| T | 5575 hCoV-19/England/NORW-F4FE4/2020\|EPI_ISL_665881\|202 | ···················· | ···················· | ····················· | ···················· | ···················· | ···················· |
| T | 5576 hCoV-19/England/NORW-F5086/2020\|EPI_ISL_665882\|202 | ···················· | ···················· | ····················· | ···················· | ···················· | ···················· |
| T | 5577 hCoV-19/England/NORW-F50A4/2020\|EPI_ISL_665883\|202 | ···················· | ···················· | ····················· | ···················· | NN·················· | ···················· |
| T | 5578 hCoV-19/England/NORW-F511A/2020\|EPI_ISL_665884\|202 | ···················· | ···················· | ····················· | ···················· | ···················· | ···················· |
| T | 5579 hCoV-19/England/NORW-F5129/2020\|EPI_ISL_665885\|202 | ···················· | ···················· | ····················· | ···················· | ···················· | ···················· |
| T | 5581 hCoV-19/England/QEUH-B7F20B/2020\|EPI_ISL_668001\|20 | ···················· | ···················· | ····················· | ···················· | ···················· | ···················· |
| T | 5582 hCoV-19/England/CAMC-B7981E/2020\|EPI_ISL_668002\|20 | ···················· | ···················· | ····················· | ···················· | ···················· | ···················· |
| T | 5583 hCoV-19/England/QEUH-B81A95/2020\|EPI_ISL_668004\|20 | ···················· | ···················· | ····················· | ···················· | ···················· | ···················· |
| T | 5584 hCoV-19/England/QEUH-B7E8EC/2020\|EPI_ISL_668005\|20 | ···················· | ···················· | ····················· | ···················· | ···················· | ···················· |
| T | 5585 hCoV-19/England/QEUH-B7EDCF/2020\|EPI_ISL_668006\|20 | ···················· | ···················· | ····················· | ···················· | ···················· | ···················· |
| T | 5586 hCoV-19/England/CAMC-B7893A/2020\|EPI_ISL_668009\|20 | ···················· | ···················· | ····················· | ···················· | ···················· | ···················· |
| T | 5587 hCoV-19/England/QEUH-B7EEAE/2020\|EPI_ISL_668013\|20 | ···················· | ·A·················· | ····················· | ···················· | ···················· | ···················· |
| T | 5588 hCoV-19/England/CAMC-B7919B/2020\|EPI_ISL_668014\|20 | ···················· | ···················· | ····················· | ···················· | ···················· | ···················· |
| T | 5589 hCoV-19/England/CAMC-B79D6A/2020\|EPI_ISL_668019\|20 | ···················· | ···················· | ····················· | ···················· | ···················· | ···················· |
| T | 5590 hCoV-19/England/CAMC-B78A91/2020\|EPI_ISL_668080\|20 | ···················· | ···················· | ····················· | ···················· | ···················· | ···················· |
| T | 5591 hCoV-19/England/QEUH-B7EE44/2020\|EPI_ISL_668082\|20 | ···················· | ···················· | ····················· | ···················· | ···················· | ···················· |
| T | 5592 hCoV-19/England/QEUH-B816CE/2020\|EPI_ISL_668084\|20 | ···················· | ···················· | ····················· | ···················· | ···················· | ···················· |
| T | 5593 hCoV-19/England/CAMC-B7A9ED/2020\|EPI_ISL_668085\|20 | ···················· | ···················· | ····················· | ···················· | ···················· | ···················· |
| T | 5594 hCoV-19/England/CAMC-B799EE/2020\|EPI_ISL_668086\|20 | ···················· | ···················· | ····················· | ···················· | ···················· | ···················· |
| T | 5595 hCoV-19/England/CAMC-B79957/2020\|EPI_ISL_668087\|20 | ···················· | ···················· | ····················· | ···················· | ···················· | ···················· |
| T | 5596 hCoV-19/England/CAMC-B79D88/2020\|EPI_ISL_668089\|20 | ···················· | ···················· | ····················· | ···················· | ···················· | ···················· |
| T | 5597 hCoV-19/England/CAMC-B7B08D/2020\|EPI_ISL_668090\|20 | ···················· | ···················· | ····················· | ···················· | ···················· | ···················· |
| T | 5598 hCoV-19/England/CAMC-B78A46/2020\|EPI_ISL_668091\|20 | ···················· | ·A·················· | ····················· | ···················· | ···················· | ··A················· |
| T | 5599 hCoV-19/England/QEUH-B81664/2020\|EPI_ISL_668093\|20 | ···················· | ···················· | ····················· | ···················· | ···················· | ···················· |
| T | 5600 hCoV-19/England/QEUH-B81673/2020\|EPI_ISL_668094\|20 | ···················· | ·A·················· | ····················· | ···················· | ···················· | ··A················· |
| T | 5601 hCoV-19/England/QEUH-B7E5B8/2020\|EPI_ISL_668095\|20 | ···················· | ···················· | ····················· | ···················· | ···················· | ···················· |
| T | 5602 hCoV-19/England/QEUH-B7EA31/2020\|EPI_ISL_668096\|20 | ···················· | ···················· | ····················· | ···················· | ···················· | ···················· |
| T | 5603 hCoV-19/England/CAMC-B79A18/2020\|EPI_ISL_668097\|20 | ···················· | ···················· | ····················· | ···················· | ···················· | ···················· |
| T | 5604 hCoV-19/England/QEUH-B7E934/2020\|EPI_ISL_668098\|20 | ···················· | ···················· | ····················· | ···················· | ···················· | ···················· |
| T | 5605 hCoV-19/Denmark/DCGC-10007/2020\|EPI_ISL_669330\|202 | ···················· | ···················· | ····················· | ···················· | ···················· | ···················· |
| T | 5606 hCoV-19/Denmark/DCGC-10008/2020\|EPI_ISL_669331\|202 | ···················· | ···················· | ····················· | ···················· | ···················· | ···················· |
| T | 5607 hCoV-19/Denmark/DCGC-10009/2020\|EPI_ISL_669332\|202 | ···················· | ···················· | ····················· | ···················· | ···················· | ···················· |
| T | 5608 hCoV-19/Denmark/DCGC-10011/2020\|EPI_ISL_669333\|202 | ···················· | ···················· | ····················· | ···················· | ···················· | ···················· |
| T | 5609 hCoV-19/Denmark/DCGC-10013/2020\|EPI_ISL_669334\|202 | ···················· | ···················· | ····················· | ···················· | ···················· | ···················· |
| T | 5610 hCoV-19/Denmark/DCGC-10014/2020\|EPI_ISL_669335\|202 | ···················· | ···················· | ····················· | ···················· | ···················· | ···················· |
| T | 5611 hCoV-19/Denmark/DCGC-10016/2020\|EPI_ISL_669336\|202 | ···················· | ···················· | ····················· | ···················· | ···················· | ···················· |
| T | 5612 hCoV-19/Denmark/DCGC-10017/2020\|EPI_ISL_669337\|202 | ···················· | ···················· | ····················· | ···················· | ···················· | ···················· |
| T | 5613 hCoV-19/Denmark/DCGC-10018/2020\|EPI_ISL_669338\|202 | ···················· | ···················· | ····················· | ···················· | ···················· | ···················· |
| T | 5614 hCoV-19/Denmark/DCGC-10019/2020\|EPI_ISL_669339\|202 | ···················· | ···················· | ····················· | ···················· | ···················· | ···················· |
| T | 5615 hCoV-19/Denmark/DCGC-10026/2020\|EPI_ISL_669344\|202 | ···················· | ···················· | ····················· | ···················· | ···················· | ···················· |
| T | 5616 hCoV-19/Denmark/DCGC-10027/2020\|EPI_ISL_669345\|202 | ···················· | ···················· | ····················· | ···················· | ···················· | ···················· |
| T | 5617 hCoV-19/Denmark/DCGC-10031/2020\|EPI_ISL_669348\|202 | ···················· | ···················· | ····················· | ···················· | ···················· | ···················· |
| T | 5618 hCoV-19/Denmark/DCGC-10032/2020\|EPI_ISL_669349\|202 | ···················· | ···················· | ····················· | ···················· | ···················· | ···················· |
| T | 5619 hCoV-19/Denmark/DCGC-10958/2020\|EPI_ISL_670310\|202 | ···················· | ···················· | ····················· | ···················· | ···················· | ···················· |
| T | 5620 hCoV-19/Denmark/DCGC-10961/2020\|EPI_ISL_670311\|202 | ···················· | ···················· | ····················· | ···················· | ······T············· | ···················· |
| T | 5621 hCoV-19/Denmark/DCGC-10962/2020\|EPI_ISL_670312\|202 | ···················· | ···················· | ····················· | ···················· | ···················· | ···················· |
| T | 5622 hCoV-19/Denmark/DCGC-10967/2020\|EPI_ISL_670313\|202 | ···················· | ···················· | ····················· | ···················· | ···················· | ···················· |
| T | 5623 hCoV-19/Denmark/DCGC-10968/2020\|EPI_ISL_670314\|202 | ···················· | ···················· | ····················· | ···················· | ···················· | ···················· |
| T | 5624 hCoV-19/Denmark/DCGC-10969/2020\|EPI_ISL_670315\|202 | ···················· | ···················· | ····················· | ···················· | ···················· | ···················· |
| T | 5625 hCoV-19/Denmark/DCGC-10970/2020\|EPI_ISL_670316\|202 | ···················· | ···················· | ····················· | ···················· | ···················· | ···················· |
| T | 5626 hCoV-19/Denmark/DCGC-10972/2020\|EPI_ISL_670317\|202 | ···················· | ···················· | ····················· | ···················· | ···················· | ···················· |
| T | 5627 hCoV-19/Denmark/DCGC-10973/2020\|EPI_ISL_670318\|202 | ···················· | ···················· | ····················· | ···················· | ···················· | ···················· |
| T | 5628 hCoV-19/Denmark/DCGC-10975/2020\|EPI_ISL_670319\|202 | ···················· | ···················· | ····················· | ···················· | ···················· | ···················· |
| T | 5629 hCoV-19/Denmark/DCGC-11110/2020\|EPI_ISL_670397\|202 | ···················· | ···················· | ····················· | ···················· | ···················· | ···················· |
| T | 5630 hCoV-19/Denmark/DCGC-11111/2020\|EPI_ISL_670398\|202 | ···················· | ···················· | ····················· | ···················· | ···················· | ···················· |
| T | 5631 hCoV-19/Denmark/DCGC-11116/2020\|EPI_ISL_670399\|202 | ···················· | ···················· | ····················· | ···················· | ···················· | ···················· |

| | | | | | | | |
|---|---|---|---|---|---|---|---|
| T | 5632 hCoV-19/USA/TX-DSHS-1448/2020\|EPI_ISL_671650\|2020- | ···················· | ············A······· | ····················· | ···················· | ···················· | ···················· |
| T | 5633 hCoV-19/Sweden/20-53380/2020\|EPI_ISL_661300\|2020-1 | ···················· | ···················· | ····················· | ···················· | ···················· | ···················· |
| T | 5634 hCoV-19/Sweden/20-53401/2020\|EPI_ISL_661301\|2020-1 | ···················· | ···················· | ····················· | ···················· | ···················· | ···················· |
| T | 5635 hCoV-19/Sweden/20-53474/2020\|EPI_ISL_661302\|2020-1 | ···················· | ···················· | ····················· | ···················· | ···················· | ···················· |
| T | 5636 hCoV-19/Sweden/20-53492/2020\|EPI_ISL_661303\|2020-1 | ···················· | ···················· | ····················· | ···················· | ···················· | ···················· |
| T | 5637 hCoV-19/India/WB-IICB-031/2020\|EPI_ISL_661304\|2020 | ···················· | ···················· | ····················· | ···················· | ···················· | ···················· |
| T | 5638 hCoV-19/India/WB-IICB-032/2020\|EPI_ISL_661305\|2020 | ···················· | ·A·················· | ····················· | ···················· | ···················· | ···················· |
| T | 5639 hCoV-19/India/WB-IICB-033/2020\|EPI_ISL_661306\|2020 | ···················· | ···················· | ····················· | ···················· | ···················· | ···················· |
| T | 5640 hCoV-19/India/WB-IICB-034/2020\|EPI_ISL_661307\|2020 | ···················· | ···················· | ····················· | ···················· | ···················· | ···················· |
| T | 5641 hCoV-19/India/WB-IICB-035/2020\|EPI_ISL_661308\|2020 | ···················· | ·A·················· | ····················· | ···················· | ···················· | ···················· |
| T | 5642 hCoV-19/India/WB-IICB-036/2020\|EPI_ISL_661309\|2020 | ···················· | ···················· | ····················· | ···················· | ···················· | ···················· |
| T | 5643 hCoV-19/India/WB-IICB-037/2020\|EPI_ISL_661310\|2020 | ···················· | ·A·················· | ····················· | ···················· | ···················· | ···················· |
| T | 5644 hCoV-19/India/WB-IICB-038/2020\|EPI_ISL_661311\|2020 | ···················· | ·A·················· | ····················· | ···················· | ···················· | ···················· |
| T | 5645 hCoV-19/England/QEUH-B67084/2020\|EPI_ISL_661314\|20 | ···················· | ···················· | ····················· | ···················· | ···················· | ···················· |
| T | 5646 hCoV-19/England/QEUH-B66EE5/2020\|EPI_ISL_661315\|20 | ···················· | ···················· | ····················· | ···················· | ···················· | ···················· |
| T | 5647 hCoV-19/England/QEUH-B66F79/2020\|EPI_ISL_661316\|20 | ···················· | ···················· | ····················· | ···················· | ···················· | ···················· |
| T | 5648 hCoV-19/England/QEUH-B683A8/2020\|EPI_ISL_661319\|20 | ···················· | ···················· | ····················· | ···················· | ···················· | ···················· |
| T | 5649 hCoV-19/England/QEUH-B66498/2020\|EPI_ISL_661320\|20 | ···················· | ···················· | ····················· | ···················· | ···················· | ···················· |
| T | 5650 hCoV-19/England/QEUH-B697E7/2020\|EPI_ISL_661355\|20 | ···················· | ···················· | ····················· | ···················· | ···················· | ···················· |
| T | 5651 hCoV-19/England/QEUH-B69820/2020\|EPI_ISL_661356\|20 | ···················· | ···················· | ····················· | ···················· | ···················· | ···················· |
| T | 5652 hCoV-19/England/QEUH-B6779E/2020\|EPI_ISL_661358\|20 | ···················· | ···················· | ····················· | ···················· | ···················· | ···················· |
| T | 5653 hCoV-19/England/QEUH-B68C61/2020\|EPI_ISL_661366\|20 | ···················· | ···················· | ····················· | ···················· | ···················· | ···················· |
| T | 5654 hCoV-19/England/QEUH-B6C5EA/2020\|EPI_ISL_661369\|20 | ···················· | ···················· | ····················· | ···················· | ···················· | ···················· |
| T | 5655 hCoV-19/England/QEUH-B6990F/2020\|EPI_ISL_661370\|20 | ···················· | ···················· | ····················· | ···················· | ······T············· | ···················· |
| T | 5656 hCoV-19/England/QEUH-B69A0C/2020\|EPI_ISL_661371\|20 | ···················· | ···················· | ····················· | ···················· | ···················· | ···················· |
| T | 5657 hCoV-19/England/QEUH-B67AA4/2020\|EPI_ISL_661372\|20 | ···················· | ···················· | ····················· | ···················· | ···················· | ···················· |
| T | 5658 hCoV-19/England/QEUH-B6646B/2020\|EPI_ISL_661373\|20 | ···················· | ·A·················· | ····················· | ···················· | ···················· | ···················· |
| T | 5659 hCoV-19/England/QEUH-B66683/2020\|EPI_ISL_661374\|20 | ···················· | ···················· | ····················· | ···················· | ···················· | ···················· |
| T | 5660 hCoV-19/England/QEUH-B67B56/2020\|EPI_ISL_661375\|20 | ···················· | ···················· | ····················· | ···················· | ···················· | ···················· |
| T | 5661 hCoV-19/England/QEUH-B693C5/2020\|EPI_ISL_661376\|20 | ···················· | ·A·················· | ····················· | ···················· | ···················· | ···················· |
| T | 5662 hCoV-19/England/QEUH-B66331/2020\|EPI_ISL_661378\|20 | ···················· | ···················· | ····················· | ···················· | ···················· | ···················· |
| T | 5663 hCoV-19/England/QEUH-B664D4/2020\|EPI_ISL_661380\|20 | ···················· | ···················· | ····················· | ···················· | ···················· | ···················· |
| T | 5664 hCoV-19/England/QEUH-B67BCF/2020\|EPI_ISL_661382\|20 | ···················· | ···················· | ····················· | ···················· | ···················· | ···················· |
| T | 5665 hCoV-19/England/QEUH-B6835D/2020\|EPI_ISL_661383\|20 | ···················· | ···················· | ····················· | ···················· | ···················· | ···················· |
| T | 5666 hCoV-19/England/QEUH-B69A93/2020\|EPI_ISL_661384\|20 | ···················· | ···················· | ····················· | ···················· | ···················· | ···················· |
| T | 5667 hCoV-19/England/QEUH-B666CF/2020\|EPI_ISL_661385\|20 | ···················· | ···················· | ····················· | ···················· | ···················· | ···················· |
| T | 5668 hCoV-19/England/QEUH-B692C8/2020\|EPI_ISL_661387\|20 | ···················· | ···················· | ····················· | ···················· | ···················· | ···················· |
| T | 5669 hCoV-19/England/QEUH-B670C0/2020\|EPI_ISL_661388\|20 | ···················· | ···················· | ····················· | ···················· | ···················· | ···················· |
| T | 5670 hCoV-19/England/QEUH-B67725/2020\|EPI_ISL_661389\|20 | ···················· | ···················· | ····················· | ···················· | ···················· | ···················· |
| T | 5671 hCoV-19/England/QEUH-B69B54/2020\|EPI_ISL_661390\|20 | ···················· | ···················· | ····················· | ···················· | ···················· | ···················· |
| T | 5672 hCoV-19/England/QEUH-B69A48/2020\|EPI_ISL_661391\|20 | ···················· | ···················· | ····················· | ···················· | ···················· | ···················· |
| T | 5673 hCoV-19/England/QEUH-B68496/2020\|EPI_ISL_661392\|20 | ···················· | ···················· | ····················· | ···················· | ···················· | ···················· |
| T | 5674 hCoV-19/England/QEUH-B688D6/2020\|EPI_ISL_661393\|20 | ···················· | ···················· | ····················· | ···················· | ···················· | ···················· |
| T | 5675 hCoV-19/England/QEUH-B692E6/2020\|EPI_ISL_661394\|20 | ···················· | ···················· | ····················· | ···················· | ···················· | ···················· |
| T | 5676 hCoV-19/England/QEUH-B6841E/2020\|EPI_ISL_661396\|20 | ···················· | ···················· | ····················· | ···················· | ···················· | ···················· |
| T | 5677 hCoV-19/England/QEUH-B69A2A/2020\|EPI_ISL_661397\|20 | ···················· | ···················· | ····················· | ···················· | ···················· | ···················· |
| T | 5678 hCoV-19/England/QEUH-B677BC/2020\|EPI_ISL_661398\|20 | ···················· | ···················· | ····················· | ···················· | ···················· | ···················· |
| T | 5679 hCoV-19/England/QEUH-B66629/2020\|EPI_ISL_661399\|20 | ···················· | ···················· | ····················· | ···················· | ···················· | ···················· |
| T | 5680 hCoV-19/England/QEUH-B6B053/2020\|EPI_ISL_662600\|20 | ···················· | ···················· | ····················· | ···················· | ···················· | ···················· |
| T | 5681 hCoV-19/England/QEUH-B6A7E6/2020\|EPI_ISL_662603\|20 | ···················· | ·A·················· | ····················· | ···················· | ···················· | ···················· |
| T | 5682 hCoV-19/England/ALDP-B7342F/2020\|EPI_ISL_662604\|20 | ···················· | ···················· | ····················· | ···················· | ···················· | ···················· |
| T | 5683 hCoV-19/England/ALDP-B73A4B/2020\|EPI_ISL_662605\|20 | ···················· | ···················· | ····················· | ···················· | ···················· | ···················· |
| T | 5684 hCoV-19/England/QEUH-B6B703/2020\|EPI_ISL_662607\|20 | ···················· | ···················· | ····················· | ···················· | ···················· | ···················· |
| T | 5685 hCoV-19/England/ALDP-B73A69/2020\|EPI_ISL_662609\|20 | ···················· | ···················· | ····················· | ···················· | ···················· | ···················· |
| T | 5686 hCoV-19/England/ALDP-B736CF/2020\|EPI_ISL_662610\|20 | ···················· | ···················· | ····················· | ···················· | ···················· | ···················· |
| T | 5687 hCoV-19/England/QEUH-B6AA83/2020\|EPI_ISL_662611\|20 | ···················· | ···················· | ····················· | ···················· | ···················· | ···················· |
| T | 5688 hCoV-19/England/QEUH-B6A3E2/2020\|EPI_ISL_662612\|20 | ···················· | ···················· | ····················· | ···················· | ···················· | ···················· |
| T | 5689 hCoV-19/England/QEUH-B6750D/2020\|EPI_ISL_662613\|20 | ···················· | ·A·················· | ····················· | ···················· | ···················· | ···················· |
| T | 5690 hCoV-19/England/ALDP-B7352C/2020\|EPI_ISL_662614\|20 | ···················· | ···················· | ····················· | ···················· | ···················· | ···················· |
| T | 5691 hCoV-19/England/QEUH-B66C18/2020\|EPI_ISL_662615\|20 | ···················· | ···················· | ····················· | ···················· | ···················· | ···················· |
| T | 5692 hCoV-19/England/ALDP-B73911/2020\|EPI_ISL_662617\|20 | ···················· | ···················· | ····················· | ···················· | ···················· | ···················· |
| T | 5693 hCoV-19/England/QEUH-B67585/2020\|EPI_ISL_662618\|20 | ···················· | ···················· | ····················· | ···················· | ···················· | ···················· |
| T | 5694 hCoV-19/England/QEUH-B6A528/2020\|EPI_ISL_662619\|20 | ···················· | ···················· | ····················· | ···················· | ···················· | ···················· |
| T | 5695 hCoV-19/England/QEUH-B6A1F7/2020\|EPI_ISL_662620\|20 | ···················· | ···················· | ····················· | ···················· | ···················· | ···················· |
| T | 5696 hCoV-19/England/ALDP-B736FC/2020\|EPI_ISL_662621\|20 | ···················· | ···················· | ····················· | ···················· | ···················· | ···················· |
| T | 5697 hCoV-19/England/ALDP-B73A0F/2020\|EPI_ISL_662622\|20 | ···················· | ···················· | ····················· | ···················· | ···················· | ···················· |
| T | 5698 hCoV-19/England/ALDP-B5C635/2020\|EPI_ISL_662623\|20 | ···················· | ···················· | ····················· | ···················· | ···················· | ···················· |
| T | 5699 hCoV-19/England/ALDP-B5BE98/2020\|EPI_ISL_662624\|20 | ···················· | ···················· | ····················· | ···················· | ···················· | ···················· |
| T | 5700 hCoV-19/England/QEUH-B6C0E9/2020\|EPI_ISL_662627\|20 | ···················· | ···················· | ····················· | ···················· | ···················· | ···················· |
| T | 5701 hCoV-19/England/ALDP-B5B8E5/2020\|EPI_ISL_662628\|20 | ···················· | ···················· | ····················· | ···················· | ···················· | ···················· |
| T | 5702 hCoV-19/England/ALDP-B5B3A8/2020\|EPI_ISL_662629\|20 | ···················· | ···················· | ····················· | ···················· | ···················· | ···················· |
| T | 5703 hCoV-19/England/ALDP-B5BDF5/2020\|EPI_ISL_662630\|20 | ···················· | ···················· | ····················· | ···················· | ···················· | ···················· |
| T | 5704 hCoV-19/England/CAMC-B51D77/2020\|EPI_ISL_662632\|20 | ···················· | ···················· | ····················· | ···················· | ······T············· | ···················· |
| T | 5705 hCoV-19/England/MILK-B54DED/2020\|EPI_ISL_662633\|20 | ···················· | ···················· | ····················· | ···················· | ···················· | ···················· |
| T | 5706 hCoV-19/England/MILK-B5A260/2020\|EPI_ISL_662634\|20 | ···················· | ···················· | ····················· | ···················· | ···················· | ···················· |
| T | 5707 hCoV-19/England/CAMC-B53470/2020\|EPI_ISL_662635\|20 | ···················· | ·A·················· | ····················· | ···················· | ···················· | ···················· |
| T | 5708 hCoV-19/England/MILK-B569E7/2020\|EPI_ISL_662636\|20 | ···················· | ·A·················· | ····················· | ···················· | ···················· | ··A················· |
| T | 5709 hCoV-19/England/MILK-B38BE5/2020\|EPI_ISL_662637\|20 | ···················· | ···················· | ····················· | ···················· | ······T············· | ···················· |
| T | 5710 hCoV-19/England/MILK-B38B7C/2020\|EPI_ISL_662638\|20 | ···················· | ·A·················· | ····················· | ···················· | ···················· | ··A················· |
| T | 5711 hCoV-19/England/MILK-B38329/2020\|EPI_ISL_662640\|20 | ···················· | ···················· | ····················· | ···················· | ···················· | ···················· |

| | | | | | | | | |
|---|---|---|---|---|---|---|---|---|
| T | 5712 hCoV-19/England/MILK-B3875A/2020\|EPI_ISL_662643\|20 | ···················· | ···················· | ····················· | ···················· | ···················· | ···················· | |
| T | 5713 hCoV-19/England/MILK-B38D85/2020\|EPI_ISL_662644\|20 | ···················· | ···················· | ····················· | ···················· | ···················· | ···················· | |
| T | 5714 hCoV-19/England/MILK-B386D5/2020\|EPI_ISL_662645\|20 | ···················· | ···················· | ····················· | ···················· | ···················· | ···················· | |
| T | 5715 hCoV-19/England/MILK-B3831A/2020\|EPI_ISL_662646\|20 | ···················· | ···················· | ····················· | ···················· | ···················· | ···················· | |
| T | 5716 hCoV-19/England/ALDP-B47D65/2020\|EPI_ISL_662647\|20 | ···················· | ···················· | ····················· | ···················· | ···················· | ···················· | |
| T | 5717 hCoV-19/England/ALDP-B48292/2020\|EPI_ISL_662648\|20 | ···················· | ···················· | ····················· | ···················· | ···················· | ···················· | |
| T | 5718 hCoV-19/England/ALDP-B482A1/2020\|EPI_ISL_662649\|20 | ···················· | ···················· | ····················· | ···················· | ···················· | ···················· | |
| T | 5719 hCoV-19/England/ALDP-B482B0/2020\|EPI_ISL_662650\|20 | ···················· | ···················· | ····················· | ···················· | ···················· | ···················· | |
| T | 5720 hCoV-19/England/CAMC-B34527/2020\|EPI_ISL_662651\|20 | ···················· | ···················· | ····················· | ···················· | ···················· | ···················· | |
| T | 5721 hCoV-19/England/CAMC-B34800/2020\|EPI_ISL_662652\|20 | ···················· | ···················· | ····················· | ···················· | ···················· | ···················· | |
| T | 5722 hCoV-19/England/QEUH-AF0CC9/2020\|EPI_ISL_662688\|20 | ···················· | ·A·················· | ····················· | ···················· | ···················· | ···················· | |
| T | 5723 hCoV-19/England/QEUH-AF0F75/2020\|EPI_ISL_662689\|20 | ···················· | ···················· | ····················· | ···················· | ···················· | ···················· | |
| T | 5724 hCoV-19/England/QEUH-AEFBB1/2020\|EPI_ISL_662698\|20 | ···················· | ···················· | ····················· | ···················· | ···················· | ···················· | |
| T | 5725 hCoV-19/England/QEUH-AEFAF0/2020\|EPI_ISL_662699\|20 | ···················· | ···················· | ····················· | ···················· | ···················· | ···················· | |
| T | 5726 hCoV-19/England/QEUH-AF5A7F/2020\|EPI_ISL_663113\|20 | ···················· | ···················· | ····················· | ···················· | ···················· | ···················· | |
| T | 5727 hCoV-19/England/QEUH-AF6768/2020\|EPI_ISL_663114\|20 | ···················· | ···················· | ····················· | ···················· | ···················· | ···················· | |
| T | 5728 hCoV-19/England/QEUH-AF51F2/2020\|EPI_ISL_663190\|20 | ···················· | ···················· | ····················· | ···················· | ···················· | ···················· | |
| T | 5729 hCoV-19/England/QEUH-AF4C10/2020\|EPI_ISL_663191\|20 | ···················· | ···················· | ····················· | ···················· | ···················· | ···················· | |
| T | 5730 hCoV-19/England/QEUH-AF4C89/2020\|EPI_ISL_663192\|20 | ···················· | ···················· | ····················· | ···················· | ···················· | ···················· | |
| T | 5731 hCoV-19/England/QEUH-AF670E/2020\|EPI_ISL_663193\|20 | ···················· | ···················· | ····················· | ···················· | ···················· | ···················· | |
| T | 5732 hCoV-19/England/QEUH-AF6610/2020\|EPI_ISL_663194\|20 | ···················· | ···················· | ····················· | ···················· | ···················· | ···················· | |
| T | 5733 hCoV-19/England/QEUH-AF4CE3/2020\|EPI_ISL_663195\|20 | ···················· | ···················· | ····················· | ···················· | ···················· | ···················· | |
| T | 5734 hCoV-19/England/QEUH-AF5DA3/2020\|EPI_ISL_663196\|20 | ···················· | ···················· | ····················· | ···················· | ···················· | ···················· | |
| T | 5735 hCoV-19/England/QEUH-AF6962/2020\|EPI_ISL_663198\|20 | ···················· | ···················· | ····················· | ···················· | ···················· | ···················· | |
| T | 5736 hCoV-19/England/QEUH-AF671D/2020\|EPI_ISL_663199\|20 | ···················· | ···················· | ····················· | ···················· | ···················· | ···················· | |
| T | 5738 hCoV-19/Australia/VIC17479/2020\|EPI_ISL_663901\|202 | ···················· | ···················· | ····················· | ···················· | ···················· | ···················· | |
| T | 5739 hCoV-19/Australia/VIC17481/2020\|EPI_ISL_663902\|202 | ···················· | ···················· | ····················· | ···················· | ···················· | ···················· | |
| T | 5740 hCoV-19/Australia/VIC17484/2020\|EPI_ISL_663903\|202 | ···················· | ···················· | ····················· | ···················· | ···················· | ···················· | |
| T | 5741 hCoV-19/Australia/VIC17497/2020\|EPI_ISL_663904\|202 | ···················· | ···················· | ····················· | ···················· | ···················· | ···················· | |
| T | 5742 hCoV-19/Australia/VIC17511/2020\|EPI_ISL_663905\|202 | ···················· | ···················· | ····················· | ···················· | ···················· | ···················· | |
| T | 5743 hCoV-19/Australia/VIC17512/2020\|EPI_ISL_663906\|202 | ···················· | ···················· | ····················· | ···················· | ···················· | ···················· | |
| T | 5744 hCoV-19/Australia/VIC17516/2020\|EPI_ISL_663907\|202 | ···················· | ···················· | ····················· | ···················· | ···················· | ···················· | |
| T | 5745 hCoV-19/Australia/VIC17517/2020\|EPI_ISL_663908\|202 | ···················· | ···················· | ····················· | ···················· | NNN················· | ···················· | 1 |
| T | 5746 hCoV-19/Australia/VIC17527/2020\|EPI_ISL_663909\|202 | ···················· | ···················· | ····················· | ···················· | ···················· | ···················· | |
| T | 5747 hCoV-19/Australia/VIC17529/2020\|EPI_ISL_663910\|202 | ···················· | ···················· | ····················· | ···················· | ···················· | ···················· | |
| T | 5748 hCoV-19/Australia/VIC17532/2020\|EPI_ISL_663911\|202 | ···················· | ···················· | ····················· | ···················· | ···················· | ···················· | |
| T | 5749 hCoV-19/Australia/VIC17534/2020\|EPI_ISL_663912\|202 | ···················· | ···················· | ····················· | ···················· | ···················· | ···················· | |
| T | 5750 hCoV-19/Australia/VIC17535/2020\|EPI_ISL_663913\|202 | ···················· | ···················· | ····················· | ···················· | ···················· | ···················· | |
| T | 5751 hCoV-19/Australia/VIC17536/2020\|EPI_ISL_663914\|202 | ···················· | ···················· | ····················· | ···················· | ···················· | ···················· | |
| T | 5752 hCoV-19/Australia/VIC17544/2020\|EPI_ISL_663915\|202 | ···················· | ···················· | ····················· | ···················· | ···················· | ···················· | |
| T | 5753 hCoV-19/Australia/VIC17545/2020\|EPI_ISL_663916\|202 | ···················· | ···················· | ····················· | ···················· | ···················· | ···················· | |
| T | 5754 hCoV-19/Australia/VIC17546/2020\|EPI_ISL_663917\|202 | ···················· | ···················· | ····················· | ···················· | ···················· | ···················· | |
| T | 5755 hCoV-19/Australia/VIC17547/2020\|EPI_ISL_663918\|202 | ···················· | ···················· | ····················· | ···················· | ···················· | ···················· | |
| T | 5756 hCoV-19/Australia/VIC17549/2020\|EPI_ISL_663919\|202 | ···················· | ···················· | ····················· | ···················· | ···················· | ···················· | |
| T | 5757 hCoV-19/Australia/VIC17551/2020\|EPI_ISL_663920\|202 | ···················· | ···················· | ····················· | ···················· | ···················· | ···················· | |
| T | 5758 hCoV-19/Australia/VIC17554/2020\|EPI_ISL_663921\|202 | ···················· | ···················· | ····················· | ···················· | ···················· | ···················· | |
| T | 5759 hCoV-19/Australia/VIC17556/2020\|EPI_ISL_663922\|202 | ···················· | ···················· | ····················· | ···················· | ···················· | ···················· | |
| T | 5760 hCoV-19/Australia/VIC17559/2020\|EPI_ISL_663923\|202 | ···················· | ···················· | ····················· | ···················· | ···················· | ···················· | |
| T | 5761 hCoV-19/Australia/VIC17564/2020\|EPI_ISL_663924\|202 | ···················· | ···················· | ····················· | ···················· | ···················· | ···················· | |
| T | 5762 hCoV-19/Australia/VIC17566/2020\|EPI_ISL_663925\|202 | ···················· | ···················· | ····················· | ···················· | ···················· | ···················· | |
| T | 5763 hCoV-19/Australia/VIC17568/2020\|EPI_ISL_663926\|202 | ···················· | ···················· | ····················· | ···················· | ···················· | ···················· | |
| T | 5764 hCoV-19/Australia/VIC17569/2020\|EPI_ISL_663927\|202 | ···················· | ···················· | ····················· | ···················· | ···················· | ···················· | |
| T | 5765 hCoV-19/Australia/VIC17570/2020\|EPI_ISL_663928\|202 | ···················· | ···················· | ····················· | ···················· | ···················· | ···················· | |
| T | 5766 hCoV-19/Australia/VIC17572/2020\|EPI_ISL_663929\|202 | ···················· | ···················· | ····················· | ···················· | ···················· | ···················· | |
| T | 5767 hCoV-19/Australia/VIC17576/2020\|EPI_ISL_663930\|202 | ···················· | ···················· | ····················· | ···················· | ···················· | ···················· | |
| T | 5768 hCoV-19/Australia/VIC17578/2020\|EPI_ISL_663931\|202 | ···················· | ···················· | ····················· | ···················· | ···················· | ···················· | |
| T | 5769 hCoV-19/Australia/VIC17582/2020\|EPI_ISL_663932\|202 | ···················· | ···················· | ····················· | ···················· | ···················· | ···················· | |
| T | 5770 hCoV-19/Australia/VIC17585/2020\|EPI_ISL_663933\|202 | ···················· | ···················· | ····················· | ···················· | ···················· | ···················· | |
| T | 5771 hCoV-19/Australia/VIC17586/2020\|EPI_ISL_663934\|202 | ···················· | ···················· | ····················· | ···················· | ···················· | ···················· | |
| T | 5772 hCoV-19/Australia/VIC17587/2020\|EPI_ISL_663935\|202 | ···················· | ···················· | ····················· | ···················· | ···················· | ···················· | |
| T | 5773 hCoV-19/Australia/VIC17588/2020\|EPI_ISL_663936\|202 | ···················· | ···················· | ····················· | ···················· | ···················· | ···················· | |
| T | 5774 hCoV-19/Australia/VIC17589/2020\|EPI_ISL_663937\|202 | ···················· | ···················· | ····················· | ···················· | ···················· | ···················· | |
| T | 5775 hCoV-19/Australia/VIC17590/2020\|EPI_ISL_663938\|202 | ···················· | ···················· | ····················· | ···················· | ···················· | ···················· | |
| T | 5776 hCoV-19/Australia/VIC17591/2020\|EPI_ISL_663939\|202 | ···················· | ···················· | ····················· | ···················· | ···················· | ···················· | |
| T | 5777 hCoV-19/Australia/VIC17593/2020\|EPI_ISL_663940\|202 | ···················· | ···················· | ····················· | ···················· | ···················· | ···················· | |
| T | 5778 hCoV-19/Australia/VIC17595/2020\|EPI_ISL_663941\|202 | ···················· | ···················· | ····················· | ···················· | ···················· | ···················· | |
| T | 5779 hCoV-19/Australia/VIC17596/2020\|EPI_ISL_663942\|202 | ···················· | ···················· | ····················· | ···················· | ···················· | ···················· | |
| T | 5780 hCoV-19/Australia/VIC17597/2020\|EPI_ISL_663943\|202 | ···················· | ···················· | ····················· | ···················· | ···················· | ···················· | |
| T | 5781 hCoV-19/Australia/VIC17599/2020\|EPI_ISL_663944\|202 | ···················· | ···················· | ····················· | ···················· | ···················· | ···················· | |
| T | 5782 hCoV-19/Australia/VIC17602/2020\|EPI_ISL_663945\|202 | ···················· | ···················· | ····················· | ···················· | ···················· | ···················· | |
| T | 5783 hCoV-19/Australia/VIC17605/2020\|EPI_ISL_663946\|202 | ···················· | ···················· | ····················· | ···················· | ···················· | ···················· | |
| T | 5784 hCoV-19/Australia/VIC17611/2020\|EPI_ISL_663947\|202 | ···················· | ···················· | ····················· | ···················· | ···················· | ···················· | |
| T | 5785 hCoV-19/Australia/VIC17613/2020\|EPI_ISL_663948\|202 | ···················· | ···················· | ····················· | ···················· | ···················· | ···················· | |
| T | 5786 hCoV-19/Australia/VIC17616/2020\|EPI_ISL_663949\|202 | ···················· | ···················· | ····················· | ···················· | ···················· | ···················· | |
| T | 5787 hCoV-19/Australia/VIC17620/2020\|EPI_ISL_663950\|202 | ···················· | ···················· | ····················· | ···················· | ···················· | ···················· | |
| T | 5788 hCoV-19/Australia/VIC17621/2020\|EPI_ISL_663951\|202 | ···················· | ···················· | ····················· | ···················· | ···················· | ···················· | |
| T | 5789 hCoV-19/Australia/VIC17622/2020\|EPI_ISL_663952\|202 | ···················· | ···················· | ····················· | ···················· | ···················· | ···················· | |
| T | 5790 hCoV-19/Australia/VIC17624/2020\|EPI_ISL_663953\|202 | ···················· | ···················· | ····················· | ···················· | ···················· | ···················· | |
| T | 5791 hCoV-19/Australia/VIC17625/2020\|EPI_ISL_663954\|202 | ···················· | ···················· | ····················· | ···················· | ···················· | ···················· | |
| T | 5792 hCoV-19/Australia/VIC17626/2020\|EPI_ISL_663955\|202 | ···················· | ···················· | ····················· | ···················· | ···················· | ···················· | |

| | | | | | | | |
|---|---|---|---|---|---|---|---|
| T | 5793 hCoV-19/Australia/VIC17629/2020\|EPI_ISL_663956\|202 | ···················· | ···················· | ···················· | ···················· | ···················· | ···················· |
| T | 5794 hCoV-19/Australia/VIC17631/2020\|EPI_ISL_663957\|202 | ···················· | ···················· | ···················· | ···················· | ···················· | ···················· |
| T | 5795 hCoV-19/Australia/VIC17633/2020\|EPI_ISL_663958\|202 | ···················· | ···················· | ···················· | ···················· | ···················· | ···················· |
| T | 5796 hCoV-19/Australia/VIC17634/2020\|EPI_ISL_663959\|202 | ···················· | ···················· | ···················· | ···················· | ···················· | ···················· |
| T | 5797 hCoV-19/Australia/VIC17636/2020\|EPI_ISL_663960\|202 | ···················· | ···················· | ···················· | ···················· | ···················· | ···················· |
| T | 5798 hCoV-19/Australia/VIC17639/2020\|EPI_ISL_663961\|202 | ···················· | ···················· | ···················· | ···················· | ···················· | ···················· |
| T | 5799 hCoV-19/Australia/VIC17641/2020\|EPI_ISL_663962\|202 | ···················· | ···················· | ···················· | ···················· | ···················· | ···················· |
| T | 5800 hCoV-19/Australia/VIC17642/2020\|EPI_ISL_663963\|202 | ···················· | ···················· | ···················· | ···················· | ···················· | ···················· |
| T | 5801 hCoV-19/Australia/VIC17645/2020\|EPI_ISL_663964\|202 | ···················· | ···················· | ···················· | ···················· | ···················· | ···················· |
| T | 5802 hCoV-19/Australia/VIC17646/2020\|EPI_ISL_663965\|202 | ···················· | ···················· | ···················· | ···················· | ···················· | ···················· |
| T | 5804 hCoV-19/Australia/VIC17648/2020\|EPI_ISL_663967\|202 | ···················· | ···················· | ···················· | ···················· | ···················· | ···················· |
| T | 5805 hCoV-19/Australia/VIC17649/2020\|EPI_ISL_663968\|202 | ···················· | ···················· | ···················· | ···················· | ···················· | ···················· |
| T | 5806 hCoV-19/Australia/VIC17650/2020\|EPI_ISL_663969\|202 | ···················· | ···················· | ···················· | ···················· | ···················· | ···················· |
| T | 5807 hCoV-19/Australia/VIC17651/2020\|EPI_ISL_663970\|202 | ···················· | ···················· | ···················· | ···················· | ···················· | ···················· |
| T | 5808 hCoV-19/Australia/VIC17652/2020\|EPI_ISL_663971\|202 | ···················· | ···················· | ···················· | ···················· | ···················· | ···················· |
| T | 5809 hCoV-19/Australia/VIC17653/2020\|EPI_ISL_663972\|202 | ···················· | ···················· | ···················· | ···················· | ···················· | ···················· |
| T | 5810 hCoV-19/Australia/VIC17654/2020\|EPI_ISL_663973\|202 | ···················· | ···················· | ···················· | ···················· | ···················· | ···················· |
| T | 5811 hCoV-19/Australia/VIC17655/2020\|EPI_ISL_663974\|202 | ···················· | ···················· | ···················· | ···················· | ···················· | ···················· |
| T | 5812 hCoV-19/Australia/VIC17656/2020\|EPI_ISL_663975\|202 | ···················· | ···················· | ···················· | ···················· | ···················· | ···················· |
| T | 5813 hCoV-19/Australia/VIC17657/2020\|EPI_ISL_663976\|202 | ···················· | ···················· | ···················· | ···················· | ···················· | ···················· |
| T | 5814 hCoV-19/Australia/VIC17658/2020\|EPI_ISL_663977\|202 | ···················· | ···················· | ···················· | ···················· | ···················· | ···················· |
| T | 5815 hCoV-19/Australia/VIC17659/2020\|EPI_ISL_663978\|202 | ···················· | ···················· | ···················· | ···················· | ···················· | ···················· |
| T | 5816 hCoV-19/Australia/VIC17660/2020\|EPI_ISL_663979\|202 | ···················· | ···················· | ···················· | ···················· | ···················· | ···················· |
| T | 5817 hCoV-19/Australia/VIC17661/2020\|EPI_ISL_663980\|202 | ···················· | ···················· | ···················· | ···················· | ···················· | ···················· |
| T | 5818 hCoV-19/Australia/VIC17662/2020\|EPI_ISL_663981\|202 | ···················· | ···················· | ···················· | ···················· | ···················· | ···················· |
| T | 5819 hCoV-19/Australia/VIC17663/2020\|EPI_ISL_663982\|202 | ···················· | ···················· | ···················· | ···················· | ···················· | ···················· |
| T | 5820 hCoV-19/Australia/VIC17664/2020\|EPI_ISL_663983\|202 | ···················· | ···················· | ···················· | ···················· | ···················· | ···················· |
| T | 5821 hCoV-19/Wales/PHWC-48BF66/2020\|EPI_ISL_665773\|2020 | ···················· | ···················· | ···················· | ···················· | ···················· | ···················· |
| T | 5822 hCoV-19/Wales/PHWC-48C26C/2020\|EPI_ISL_665774\|2020 | ···················· | ···················· | ···················· | ···················· | ···················· | ···················· |
| T | 5823 hCoV-19/Wales/PHWC-48AF94/2020\|EPI_ISL_666180\|2020 | ···················· | ···················· | ···················· | ···················· | ···················· | ···················· |
| T | 5824 hCoV-19/Wales/PHWC-48AFA3/2020\|EPI_ISL_666181\|2020 | ···················· | ···················· | ···················· | ···················· | ···················· | ···················· |
| T | 5825 hCoV-19/Wales/PHWC-48AFEF/2020\|EPI_ISL_666182\|2020 | ···················· | ···················· | ···················· | ···················· | ···················· | ···················· |
| T | 5826 hCoV-19/Wales/PHWC-48B009/2020\|EPI_ISL_666183\|2020 | ···················· | ···················· | ···················· | ···················· | ···················· | ···················· |
| T | 5827 hCoV-19/Wales/PHWC-48B018/2020\|EPI_ISL_666184\|2020 | ···················· | ···················· | ···················· | ···················· | ···················· | ···················· |
| T | 5829 hCoV-19/Wales/PHWC-48B045/2020\|EPI_ISL_666186\|2020 | ···················· | ···················· | ···················· | ···················· | ···················· | ···················· |
| T | 5830 hCoV-19/Wales/PHWC-48B054/2020\|EPI_ISL_666187\|2020 | ···················· | ···················· | ···················· | ···················· | ···················· | ···················· |
| T | 5831 hCoV-19/Wales/PHWC-48B063/2020\|EPI_ISL_666188\|2020 | ···················· | ···················· | ···················· | ···················· | ···················· | ···················· |
| T | 5832 hCoV-19/Wales/PHWC-48B081/2020\|EPI_ISL_666189\|2020 | ···················· | ···················· | ···················· | ···················· | ···················· | ···················· |
| T | 5833 hCoV-19/Wales/PHWC-48B090/2020\|EPI_ISL_666190\|2020 | ···················· | ···················· | ···················· | ···················· | ···················· | ···················· |
| T | 5835 hCoV-19/Wales/PHWC-48B0BE/2020\|EPI_ISL_666192\|2020 | ···················· | ···················· | ···················· | ···················· | ···················· | ···················· |
| T | 5836 hCoV-19/Wales/PHWC-48B0EB/2020\|EPI_ISL_666193\|2020 | ···················· | ···················· | ···················· | ···················· | ···················· | ···················· |
| T | 5837 hCoV-19/Wales/PHWC-48B0FA/2020\|EPI_ISL_666194\|2020 | ···················· | ···················· | ···················· | ···················· | ···················· | ···················· |
| T | 5839 hCoV-19/Wales/PHWC-48B124/2020\|EPI_ISL_666196\|2020 | ···················· | ···················· | ···················· | ···················· | ···················· | ···················· |
| T | 5840 hCoV-19/Wales/PHWC-48B142/2020\|EPI_ISL_666197\|2020 | ···················· | ···················· | ···················· | ···················· | ···················· | ···················· |
| T | 5841 hCoV-19/Wales/PHWC-48B151/2020\|EPI_ISL_666198\|2020 | ···················· | ···················· | ···················· | ···················· | ···················· | ···················· |
| T | 5842 hCoV-19/Wales/PHWC-48B160/2020\|EPI_ISL_666199\|2020 | ···················· | ···················· | ···················· | ···················· | ···················· | ···················· |
| T | 5843 hCoV-19/Denmark/DCGC-9968/2020\|EPI_ISL_669300\|2020 | ···················· | ·A·················· | ···················· | ···················· | ···················· | ···················· |
| T | 5844 hCoV-19/Denmark/DCGC-9970/2020\|EPI_ISL_669301\|2020 | ···················· | ···················· | ···················· | ···················· | ···················· | ···················· |
| T | 5845 hCoV-19/Denmark/DCGC-9971/2020\|EPI_ISL_669302\|2020 | ···················· | ···················· | ···················· | ···················· | ···················· | ···················· |
| T | 5846 hCoV-19/Denmark/DCGC-9972/2020\|EPI_ISL_669303\|2020 | ···················· | ···················· | ···················· | ···················· | ···················· | ···················· |
| T | 5847 hCoV-19/Denmark/DCGC-9975/2020\|EPI_ISL_669304\|2020 | ···················· | ···················· | ···················· | ···················· | ···················· | ···················· |
| T | 5848 hCoV-19/Denmark/DCGC-9976/2020\|EPI_ISL_669305\|2020 | ···················· | ···················· | ···················· | ···················· | ···················· | ···················· |
| T | 5849 hCoV-19/Denmark/DCGC-9979/2020\|EPI_ISL_669306\|2020 | ···················· | ···················· | ···················· | ···················· | ···················· | ···················· |
| T | 5850 hCoV-19/Denmark/DCGC-9980/2020\|EPI_ISL_669307\|2020 | ···················· | ···················· | ···················· | ···················· | ···················· | ···················· |
| T | 5851 hCoV-19/Denmark/DCGC-9981/2020\|EPI_ISL_669308\|2020 | ···················· | ···················· | ···················· | ···················· | ···················· | ···················· |
| T | 5852 hCoV-19/Denmark/DCGC-9982/2020\|EPI_ISL_669309\|2020 | ···················· | ···················· | ···················· | ···················· | ···················· | ···················· |
| T | 5853 hCoV-19/Denmark/DCGC-9983/2020\|EPI_ISL_669310\|2020 | ···················· | ·········A·········· | ···················· | ···················· | ···················· | ···················· |
| T | 5854 hCoV-19/Denmark/DCGC-9986/2020\|EPI_ISL_669311\|2020 | ···················· | ···················· | ···················· | ···················· | ···················· | ···················· |
| T | 5855 hCoV-19/Denmark/DCGC-9987/2020\|EPI_ISL_669312\|2020 | ···················· | ···················· | ···················· | ···················· | ···················· | ···················· |
| T | 5856 hCoV-19/Denmark/DCGC-9989/2020\|EPI_ISL_669313\|2020 | ···················· | ···················· | ···················· | ···················· | ···················· | ···················· |
| T | 5857 hCoV-19/Denmark/DCGC-9990/2020\|EPI_ISL_669314\|2020 | ···················· | ···················· | ···················· | ···················· | ···················· | ···················· |
| T | 5858 hCoV-19/Denmark/DCGC-9991/2020\|EPI_ISL_669315\|2020 | ···················· | ···················· | ···················· | ···················· | ···················· | ···················· |
| T | 5859 hCoV-19/Denmark/DCGC-9992/2020\|EPI_ISL_669316\|2020 | ···················· | ···················· | ···················· | ···················· | ···················· | ···················· |
| T | 5860 hCoV-19/Denmark/DCGC-9993/2020\|EPI_ISL_669317\|2020 | ···················· | ···················· | ···················· | ···················· | ···················· | ···················· |
| T | 5861 hCoV-19/Denmark/DCGC-9994/2020\|EPI_ISL_669318\|2020 | ···················· | ···················· | ···················· | ···················· | ······T············· | ···················· |
| T | 5862 hCoV-19/Denmark/DCGC-9995/2020\|EPI_ISL_669319\|2020 | ···················· | ·A·················· | ···················· | ···················· | ···················· | ···················· |
| T | 5863 hCoV-19/Denmark/DCGC-9996/2020\|EPI_ISL_669320\|2020 | ···················· | ·A·················· | ···················· | ···················· | ···················· | ···················· |
| T | 5864 hCoV-19/Denmark/DCGC-9998/2020\|EPI_ISL_669321\|2020 | ···················· | ···················· | ···················· | ···················· | ···················· | ···················· |
| T | 5865 hCoV-19/Denmark/DCGC-9999/2020\|EPI_ISL_669322\|2020 | ···················· | ···················· | ···················· | ···················· | ···················· | ···················· |
| T | 5866 hCoV-19/Denmark/DCGC-10000/2020\|EPI_ISL_669323\|202 | ···················· | ···················· | ···················· | ···················· | ···················· | ···················· |
| T | 5867 hCoV-19/Denmark/DCGC-10001/2020\|EPI_ISL_669324\|202 | ···················· | ···················· | ···················· | ···················· | ···················· | ···················· |
| T | 5868 hCoV-19/Denmark/DCGC-10002/2020\|EPI_ISL_669325\|202 | ···················· | ···················· | ···················· | ···················· | ···················· | ···················· |
| T | 5869 hCoV-19/Denmark/DCGC-10003/2020\|EPI_ISL_669326\|202 | ···················· | ···················· | ···················· | ···················· | ···················· | ···················· |
| T | 5870 hCoV-19/Denmark/DCGC-10004/2020\|EPI_ISL_669327\|202 | ···················· | ···················· | ···················· | ···················· | ···················· | ···················· |
| T | 5871 hCoV-19/Denmark/DCGC-10005/2020\|EPI_ISL_669328\|202 | ···················· | ···················· | ···················· | ···················· | ···················· | ···················· |
| T | 5872 hCoV-19/Denmark/DCGC-10006/2020\|EPI_ISL_669329\|202 | ···················· | ···················· | ···················· | ···················· | ···················· | ···················· |
| T | 5873 hCoV-19/Denmark/DCGC-10943/2020\|EPI_ISL_670300\|202 | ···················· | ·A·················· | ···················· | ···················· | ···················· | ···A················ |
| T | 5874 hCoV-19/Denmark/DCGC-10944/2020\|EPI_ISL_670301\|202 | ···················· | ···················· | ···················· | ···················· | ···················· | ···················· |
| T | 5875 hCoV-19/Denmark/DCGC-10945/2020\|EPI_ISL_670302\|202 | ···················· | ···················· | ···················· | ···················· | ···················· | ···················· |
| T | 5876 hCoV-19/Denmark/DCGC-10947/2020\|EPI_ISL_670303\|202 | ···················· | ···················· | ···················· | ···················· | ···················· | ···················· |

```
T    5877 hCoV-19/Denmark/DCGC-10948/2020|EPI_ISL_670304|202    ····················  ·A··················  ····················  ····················  ····················  ··A·················
T    5878 hCoV-19/Denmark/DCGC-10951/2020|EPI_ISL_670305|202    ····················  ····················  ····················  ····················  ····················  ····················
T    5879 hCoV-19/Denmark/DCGC-10952/2020|EPI_ISL_670306|202    ····················  ····················  ····················  ····················  ····················  ····················
T    5880 hCoV-19/Denmark/DCGC-10953/2020|EPI_ISL_670307|202    ····················  ····················  ····················  ····················  ····················  ····················
T    5881 hCoV-19/Denmark/DCGC-10955/2020|EPI_ISL_670308|202    ····················  ····················  ····················  ····················  ····················  ····················
T    5882 hCoV-19/Denmark/DCGC-10956/2020|EPI_ISL_670309|202    ····················  ····················  ····················  ····················  ····················  ····················
T    5883 hCoV-19/Canada/ON-PHL-20-02344/2020|EPI_ISL_671600    ····················  ···A················  ····················  ····················  ····················  ····················
T    5884 hCoV-19/Canada/ON-PHL-20-02333/2020|EPI_ISL_671601    ····················  ····················  ····················  ····················  ····················  ····················
T    5885 hCoV-19/Canada/ON-PHL-20-02452/2020|EPI_ISL_671602    ····················  ···A················  ····················  ····················  ····················  ····················
T    5886 hCoV-19/Canada/ON-PHL-20-02482/2020|EPI_ISL_671603    ····················  ····················  ····················  ····················  ····················  ····················
T    5887 hCoV-19/Canada/ON-PHL-20-02074/2020|EPI_ISL_671605    ····················  ····················  ····················  ····················  ····················  ····················
T    5888 hCoV-19/Canada/ON-PHL-20-02415/2020|EPI_ISL_671606    ····················  ····················  ····················  ····················  ····················  ····················
T    5889 hCoV-19/Sweden/20-53240/2020|EPI_ISL_661289|2020-1    ····················  ····················  ····················  ····················  ·····T··············  ····················
T    5890 hCoV-19/Sweden/20-53262/2020|EPI_ISL_661290|2020-1    ····················  ····················  ····················  ····················  ····················  ····················
T    5891 hCoV-19/Sweden/20-53267/2020|EPI_ISL_661291|2020-1    ····················  ····················  ····················  ····················  ·····T··············  ····················
T    5892 hCoV-19/Sweden/20-53277/2020|EPI_ISL_661292|2020-1    ····················  ····················  ····················  ····················  ·····T··············  ····················
T    5893 hCoV-19/Sweden/20-53288/2020|EPI_ISL_661293|2020-1    ····················  ····················  ····················  ····················  ····················  ····················
T    5894 hCoV-19/Sweden/20-53290/2020|EPI_ISL_661294|2020-1    ····················  ····················  ····················  ····················  ····················  ····················
T    5895 hCoV-19/Sweden/20-53291/2020|EPI_ISL_661295|2020-1    ····················  ····················  ····················  ····················  ·····T··············  ····················
T    5896 hCoV-19/Sweden/20-53292/2020|EPI_ISL_661296|2020-1    ····················  ····················  ····················  ····················  ·····T··············  ····················
T    5897 hCoV-19/Sweden/20-53294/2020|EPI_ISL_661297|2020-1    ····················  ····················  ····················  ····················  ····················  ····················
T    5898 hCoV-19/Sweden/20-53295/2020|EPI_ISL_661298|2020-1    ····················  ····················  ····················  ····················  ·····T··············  ····················
T    5899 hCoV-19/Sweden/20-53330/2020|EPI_ISL_661299|2020-1    ····················  ····················  ····················  ····················  ·····T··············  ····················
T    5900 hCoV-19/England/QEUH-B698B7/2020|EPI_ISL_661322|20    ····················  ····················  ····················  ····················  ····················  ····················
T    5901 hCoV-19/England/QEUH-B69802/2020|EPI_ISL_661323|20    ····················  ····················  ····················  ····················  ····················  ····················
T    5902 hCoV-19/England/QEUH-B66692/2020|EPI_ISL_661324|20    ····················  ····················  ····················  ····················  ····················  ····················
T    5903 hCoV-19/England/QEUH-B6845A/2020|EPI_ISL_661325|20    ····················  ····················  ····················  ····················  ····················  ····················
T    5904 hCoV-19/England/QEUH-B6988A/2020|EPI_ISL_661326|20    ····················  ····················  ····················  ····················  ····················  ····················
T    5905 hCoV-19/England/QEUH-B689D3/2020|EPI_ISL_661327|20    ····················  ····················  ····················  ····················  ····················  ····················
T    5906 hCoV-19/England/QEUH-B6C5DB/2020|EPI_ISL_661328|20    ····················  ····················  ····················  ····················  ····················  ····················
T    5907 hCoV-19/England/QEUH-B6843C/2020|EPI_ISL_661329|20    ····················  ·A··················  ····················  ····················  ····················  ····················
T    5908 hCoV-19/England/QEUH-B6647A/2020|EPI_ISL_661330|20    ····················  ····················  ····················  ····················  ····················  ····················
T    5909 hCoV-19/England/QEUH-B67998/2020|EPI_ISL_661331|20    ····················  ····················  ····················  ····················  ····················  ····················
T    5910 hCoV-19/England/QEUH-B698D5/2020|EPI_ISL_661332|20    ····················  ····················  ····················  ····················  ····················  ····················
T    5911 hCoV-19/England/QEUH-B663D7/2020|EPI_ISL_661333|20    ····················  ····················  ····················  ····················  ····················  ····················
T    5912 hCoV-19/England/QEUH-B6644D/2020|EPI_ISL_661335|20    ····················  ····················  ····················  ····················  ····················  ····················
T    5913 hCoV-19/England/QEUH-B6650E/2020|EPI_ISL_661337|20    ····················  ····················  ····················  ····················  ····················  ····················
T    5914 hCoV-19/England/QEUH-B6A2E5/2020|EPI_ISL_661338|20    ····················  ····················  ····················  ····················  ····················  ····················
T    5915 hCoV-19/England/QEUH-B66E30/2020|EPI_ISL_661339|20    ····················  ····················  ····················  ····················  ····················  ····················
T    5916 hCoV-19/England/QEUH-B69A39/2020|EPI_ISL_661340|20    ····················  ····················  ····················  ····················  ····················  ····················
T    5917 hCoV-19/England/QEUH-B692F5/2020|EPI_ISL_661341|20    ····················  ····················  ····················  ····················  ····················  ····················
T    5918 hCoV-19/England/QEUH-B66577/2020|EPI_ISL_661342|20    ····················  ····················  ····················  ····················  ····················  ····················
T    5919 hCoV-19/England/QEUH-B6993C/2020|EPI_ISL_661343|20    ····················  ····················  ····················  ····················  ····················  ····················
T    5920 hCoV-19/England/QEUH-B682BA/2020|EPI_ISL_661344|20    ····················  ····················  ····················  ····················  ····················  ····················
T    5921 hCoV-19/England/QEUH-B68320/2020|EPI_ISL_661346|20    ····················  ····················  ····················  ····················  ····················  ····················
T    5922 hCoV-19/England/QEUH-B66FB5/2020|EPI_ISL_661347|20    ····················  ·A··················  ····················  ····················  ····················  ····················
T    5923 hCoV-19/England/QEUH-B662CB/2020|EPI_ISL_661348|20    ····················  ····················  ····················  ····················  ····················  ····················
T    5924 hCoV-19/England/QEUH-B67039/2020|EPI_ISL_661349|20    ····················  ····················  ····················  ····················  ····················  ····················
T    5925 hCoV-19/England/QEUH-B67057/2020|EPI_ISL_661351|20    ····················  ····················  ····················  ····················  ····················  ····················
T    5926 hCoV-19/England/QEUH-B684A5/2020|EPI_ISL_661352|20    ····················  ····················  ····················  ····················  ····················  ····················
T    5927 hCoV-19/England/QEUH-B677E9/2020|EPI_ISL_661354|20    ····················  ····················  ····················  ····················  ····················  ····················
T    5928 hCoV-19/England/QEUH-B69398/2020|EPI_ISL_661359|20    ····················  ·A··················  ····················  ····················  ····················  ····················
T    5929 hCoV-19/England/QEUH-B676BF/2020|EPI_ISL_661360|20    ····················  ····················  ····················  ····················  ····················  ····················
T    5930 hCoV-19/England/QEUH-B6983F/2020|EPI_ISL_661361|20    ····················  ····················  ····················  ····················  ····················  ····················
T    5931 hCoV-19/England/QEUH-B67691/2020|EPI_ISL_661363|20    ····················  ····················  ····················  ····················  ····················  ····················
T    5932 hCoV-19/England/QEUH-B6C641/2020|EPI_ISL_661364|20    ····················  ····················  ····················  ····················  ····················  ····················
T    5933 hCoV-19/England/QEUH-B692AA/2020|EPI_ISL_661365|20    ····················  ····················  ····················  ····················  ····················  ····················
T    5934 hCoV-19/England/QEUH-AF00FA/2020|EPI_ISL_662655|20    ····················  ····················  ····················  ····················  ····················  ····················
T    5935 hCoV-19/England/QEUH-AF2605/2020|EPI_ISL_662656|20    ····················  ····················  ····················  ····················  ····················  ····················
T    5936 hCoV-19/England/QEUH-AF0063/2020|EPI_ISL_662660|20    ····················  ····················  ····················  ····················  ····················  ····················
T    5937 hCoV-19/England/QEUH-AF1044/2020|EPI_ISL_662661|20    ····················  ····················  ····················  ····················  ····················  ····················
T    5938 hCoV-19/England/QEUH-AF280F/2020|EPI_ISL_662662|20    ····················  ····················  ····················  ····················  ····················  ····················
T    5939 hCoV-19/England/QEUH-AF28B4/2020|EPI_ISL_662663|20    ····················  ····················  ····················  ····················  ····················  ····················
T    5940 hCoV-19/England/QEUH-AF259F/2020|EPI_ISL_662668|20    ····················  ····················  ····················  ····················  ····················  ····················
T    5941 hCoV-19/England/QEUH-AF28D2/2020|EPI_ISL_662675|20    ····················  ····················  ····················  ····················  ····················  ····················
T    5942 hCoV-19/England/QEUH-AEFA87/2020|EPI_ISL_662676|20    ····················  ····················  ····················  ····················  ····················  ····················
T    5943 hCoV-19/England/QEUH-AF27B7/2020|EPI_ISL_662677|20    ····················  ····················  ····················  ····················  ····················  ····················
T    5944 hCoV-19/England/QEUH-AF0FFD/2020|EPI_ISL_662680|20    ····················  ····················  ····················  ····················  ····················  ····················
T    5945 hCoV-19/England/QEUH-AEFC81/2020|EPI_ISL_662681|20    ····················  ····················  ····················  ····················  ····················  ····················
T    5946 hCoV-19/England/QEUH-AF0036/2020|EPI_ISL_662682|20    ····················  ····················  ····················  ····················  ····················  ····················
T    5947 hCoV-19/England/QEUH-AEFD15/2020|EPI_ISL_662684|20    ····················  ····················  ····················  ····················  ·····T··············  ····················
T    5948 hCoV-19/England/QEUH-AF04FE/2020|EPI_ISL_662690|20    ····················  ····················  ····················  ····················  ····················  ····················
T    5949 hCoV-19/England/QEUH-AF46D6/2020|EPI_ISL_662691|20    ····················  ····················  ····················  ····················  ····················  ····················
T    5950 hCoV-19/England/QEUH-AF00CD/2020|EPI_ISL_662692|20    ····················  ····················  ····················  ····················  ····················  ····················
T    5951 hCoV-19/England/QEUH-AF46C7/2020|EPI_ISL_662695|20    ····················  ····················  ····················  ····················  ····················  ····················
T    5952 hCoV-19/Australia/VIC17665/2020|EPI_ISL_663984|202    ····················  ····················  ····················  ····················  ····················  ····················
T    5953 hCoV-19/Australia/VIC17666/2020|EPI_ISL_663985|202    ····················  ····················  ····················  ····················  ····················  ····················
T    5955 hCoV-19/Australia/VIC17669/2020|EPI_ISL_663987|202    ····················  ····················  ····················  ····················  ····················  ····················
T    5956 hCoV-19/Australia/VIC17670/2020|EPI_ISL_663988|202    ····················  ····················  ····················  ····················  ····················  ····················
T    5957 hCoV-19/Australia/VIC17671/2020|EPI_ISL_663989|202    ····················  ····················  ····················  ····················  ····················  ····················
```

| | | | | | | | | |
|---|---|---|---|---|---|---|---|---|
| T | 5958 hCoV-19/Australia/VIC17672/2020\|EPI_ISL_663990\|202 | ···················· | ···················· | ···················· | ···················· | ···················· | ···················· | |
| T | 5959 hCoV-19/Australia/VIC17675/2020\|EPI_ISL_663991\|202 | ···················· | ···················· | ···················· | ···················· | ···················· | ···················· | |
| T | 5960 hCoV-19/Australia/VIC17677/2020\|EPI_ISL_663992\|202 | ···················· | ···················· | ···················· | ···················· | ···················· | ···················· | |
| T | 5961 hCoV-19/Australia/VIC17678/2020\|EPI_ISL_663993\|202 | ···················· | ···················· | ···················· | ···················· | ···················· | ···················· | |
| T | 5963 hCoV-19/Australia/VIC17681/2020\|EPI_ISL_663995\|202 | ···················· | ···················· | ···················· | ···················· | ···················· | ···················· | |
| T | 5965 hCoV-19/Australia/VIC17686/2020\|EPI_ISL_663997\|202 | ···················· | ···················· | ···················· | ···················· | ···················· | ···················· | |
| T | 5966 hCoV-19/Australia/VIC17689/2020\|EPI_ISL_663998\|202 | ···················· | ···················· | ···················· | ···················· | ···················· | ···················· | |
| T | 5967 hCoV-19/Australia/VIC17692/2020\|EPI_ISL_663999\|202 | ···················· | ···················· | ···················· | ···················· | ···················· | ···················· | |
| T | 5968 hCoV-19/Scotland/QEUH-AA06AC/2020\|EPI_ISL_666100\|2 | ···················· | ···················· | ···················· | ···················· | ···················· | ···················· | |
| T | 5969 hCoV-19/Scotland/QEUH-AF0D20/2020\|EPI_ISL_666101\|2 | ···················· | ···················· | ···················· | ···················· | ···················· | ···················· | |
| T | 5970 hCoV-19/Scotland/QEUH-AF1132/2020\|EPI_ISL_666102\|2 | ···················· | ···················· | ···················· | ···················· | ···················· | ···················· | |
| T | 5971 hCoV-19/Scotland/QEUH-AF1484/2020\|EPI_ISL_666105\|2 | ···················· | ···················· | ···················· | ···················· | ···················· | ···················· | |
| T | 5972 hCoV-19/Wales/ALDP-B1CA26/2020\|EPI_ISL_666107\|2020 | ···················· | ···················· | ···················· | ···················· | ···················· | ···················· | |
| T | 5973 hCoV-19/Wales/ALDP-B1CACC/2020\|EPI_ISL_666108\|2020 | ···················· | ···················· | ···················· | ···················· | ···················· | ···················· | |
| T | 5974 hCoV-19/Wales/ALDP-B1CB32/2020\|EPI_ISL_666109\|2020 | ···················· | ···················· | ···················· | ···················· | ···················· | ···················· | |
| T | 5975 hCoV-19/Wales/PHWC-48A9D2/2020\|EPI_ISL_666110\|2020 | ···················· | ···················· | ···················· | ···················· | ···················· | ···················· | |
| T | 5976 hCoV-19/Wales/PHWC-48AA0C/2020\|EPI_ISL_666111\|2020 | ···················· | ···················· | ···················· | ···················· | ···················· | ···················· | |
| T | 5977 hCoV-19/Wales/PHWC-48AA1B/2020\|EPI_ISL_666112\|2020 | ···················· | ···················· | ···················· | ···················· | ···················· | ···················· | |
| T | 5978 hCoV-19/Wales/PHWC-48AA2A/2020\|EPI_ISL_666113\|2020 | ···················· | ···················· | ···················· | ···················· | ···················· | ···················· | |
| T | 5979 hCoV-19/Wales/PHWC-48AA39/2020\|EPI_ISL_666114\|2020 | ···················· | ···················· | ···················· | ···················· | ···················· | ···················· | |
| T | 5980 hCoV-19/Wales/PHWC-48AA57/2020\|EPI_ISL_666115\|2020 | ···················· | ···················· | ···················· | ···················· | ···················· | ···················· | |
| T | 5981 hCoV-19/Wales/PHWC-48AA84/2020\|EPI_ISL_666116\|2020 | ···················· | ···················· | ···················· | ···················· | ···················· | ···················· | |
| T | 5982 hCoV-19/Wales/PHWC-48AA93/2020\|EPI_ISL_666117\|2020 | ···················· | ···················· | ···················· | ···················· | ···················· | ···················· | |
| T | 5983 hCoV-19/Wales/PHWC-48AAB1/2020\|EPI_ISL_666118\|2020 | ···················· | ···················· | ···················· | ···················· | ···················· | ···················· | |
| T | 5984 hCoV-19/Wales/PHWC-48AAC0/2020\|EPI_ISL_666119\|2020 | ···················· | ···················· | ···················· | ···················· | ···················· | ···················· | |
| T | 5985 hCoV-19/Wales/PHWC-48AADF/2020\|EPI_ISL_666120\|2020 | ···················· | ···················· | ···················· | ···················· | ···················· | ···················· | |
| T | 5986 hCoV-19/Wales/PHWC-48AAFD/2020\|EPI_ISL_666121\|2020 | ···················· | ···················· | ···················· | ···················· | ···················· | ···················· | |
| T | 5987 hCoV-19/Wales/PHWC-48AB09/2020\|EPI_ISL_666122\|2020 | ···················· | ···················· | ···················· | ···················· | ···················· | ···················· | |
| T | 5988 hCoV-19/Wales/PHWC-48AB18/2020\|EPI_ISL_666123\|2020 | ···················· | ···················· | ···················· | ···················· | ···················· | ···················· | |
| T | 5989 hCoV-19/Wales/PHWC-48AB45/2020\|EPI_ISL_666124\|2020 | ···················· | ···················· | ···················· | ···················· | ···················· | ···················· | |
| T | 5990 hCoV-19/Wales/PHWC-48AB54/2020\|EPI_ISL_666125\|2020 | ···················· | ···················· | ···················· | ···················· | ···················· | ···················· | |
| T | 5991 hCoV-19/Wales/PHWC-48AB63/2020\|EPI_ISL_666126\|2020 | ···················· | ···················· | ···················· | ···················· | ···················· | ···················· | |
| T | 5992 hCoV-19/Wales/PHWC-48AB81/2020\|EPI_ISL_666127\|2020 | ···················· | ···················· | ···················· | ···················· | ···················· | ···················· | |
| T | 5993 hCoV-19/Wales/PHWC-48AB90/2020\|EPI_ISL_666128\|2020 | ···················· | ···················· | ···················· | ···················· | ···················· | ···················· | |
| T | 5994 hCoV-19/Wales/PHWC-48ABAF/2020\|EPI_ISL_666129\|2020 | ···················· | ···················· | ···················· | ···················· | ···················· | ···················· | |
| T | 5995 hCoV-19/Wales/PHWC-48ABBE/2020\|EPI_ISL_666130\|2020 | ···················· | ···················· | ···················· | ···················· | ···················· | ···················· | |
| T | 5996 hCoV-19/Wales/PHWC-48ABCD/2020\|EPI_ISL_666131\|2020 | ···················· | ···················· | ···················· | ···················· | ···················· | ···················· | |
| T | 5997 hCoV-19/Wales/PHWC-48ABDC/2020\|EPI_ISL_666132\|2020 | ···················· | ···················· | ···················· | ···················· | ···················· | ···················· | |
| T | 5998 hCoV-19/Wales/PHWC-48ABEB/2020\|EPI_ISL_666133\|2020 | ···················· | ···················· | ···················· | ···················· | ···················· | ···················· | |
| T | 5999 hCoV-19/Wales/PHWC-48ABFA/2020\|EPI_ISL_666134\|2020 | ···················· | ···················· | ···················· | ···················· | ···················· | ···················· | |
| T | 6001 hCoV-19/Wales/PHWC-48AC15/2020\|EPI_ISL_666136\|2020 | ···················· | ···················· | ···················· | ···················· | ···················· | ···················· | |
| T | 6002 hCoV-19/Wales/PHWC-48AC24/2020\|EPI_ISL_666137\|2020 | ···················· | ···················· | ···················· | ···················· | ···················· | ···················· | |
| T | 6003 hCoV-19/Wales/PHWC-48AC42/2020\|EPI_ISL_666138\|2020 | ···················· | ···················· | ···················· | ···················· | ···················· | ···················· | |
| T | 6004 hCoV-19/Wales/PHWC-48AC51/2020\|EPI_ISL_666139\|2020 | ···················· | ···················· | ···················· | ···················· | ···················· | ···················· | |
| T | 6005 hCoV-19/Wales/PHWC-48AC60/2020\|EPI_ISL_666140\|2020 | ···················· | ···················· | ···················· | ···················· | ···················· | ···················· | |
| T | 6006 hCoV-19/Wales/PHWC-48AC7F/2020\|EPI_ISL_666141\|2020 | ···················· | ···················· | ···················· | ···················· | ··A················· | ···················· | |
| T | 6007 hCoV-19/Wales/PHWC-48AC9D/2020\|EPI_ISL_666142\|2020 | ···················· | ···················· | ···················· | ···················· | ···················· | ···················· | |
| T | 6009 hCoV-19/Wales/PHWC-48ACBB/2020\|EPI_ISL_666144\|2020 | ···················· | ···················· | ···················· | ···················· | ···················· | ···················· | |
| T | 6010 hCoV-19/Wales/PHWC-48ACCA/2020\|EPI_ISL_666145\|2020 | ···················· | ···················· | ···················· | ···················· | ···················· | ···················· | |
| T | 6011 hCoV-19/Wales/PHWC-48ACD9/2020\|EPI_ISL_666146\|2020 | ···················· | ···················· | ···················· | ···················· | ···················· | ···················· | |
| T | 6012 hCoV-19/Wales/PHWC-48ACE8/2020\|EPI_ISL_666147\|2020 | ···················· | ···················· | ···················· | ···················· | ···················· | ···················· | |
| T | 6013 hCoV-19/Wales/PHWC-48ACF7/2020\|EPI_ISL_666148\|2020 | ···················· | ···················· | ···················· | ···················· | ···················· | ···················· | |
| T | 6014 hCoV-19/Wales/PHWC-48AD12/2020\|EPI_ISL_666149\|2020 | ···················· | ···················· | ···················· | ···················· | ···················· | ···················· | |
| T | 6015 hCoV-19/Wales/PHWC-48AD21/2020\|EPI_ISL_666150\|2020 | ···················· | ···················· | ···················· | ···················· | ···················· | ···················· | |
| T | 6016 hCoV-19/Wales/PHWC-48AD30/2020\|EPI_ISL_666151\|2020 | ···················· | ···················· | ···················· | ···················· | ···················· | ···················· | |
| T | 6017 hCoV-19/Wales/PHWC-48AD4F/2020\|EPI_ISL_666152\|2020 | ···················· | ···················· | ···················· | ···················· | ···················· | ···················· | |
| T | 6019 hCoV-19/Wales/PHWC-48AD8B/2020\|EPI_ISL_666154\|2020 | ···················· | ···················· | ···················· | ···················· | ···················· | ···················· | |
| T | 6021 hCoV-19/Wales/PHWC-48ADA9/2020\|EPI_ISL_666156\|2020 | ···················· | ···················· | ···················· | ···················· | ···················· | ···················· | |
| T | 6022 hCoV-19/Wales/PHWC-48ADB8/2020\|EPI_ISL_666157\|2020 | ···················· | ···················· | ···················· | ···················· | ···················· | ···················· | |
| T | 6023 hCoV-19/Wales/PHWC-48ADC7/2020\|EPI_ISL_666158\|2020 | ···················· | ···················· | ···················· | ···················· | ···················· | ···················· | |
| T | 6024 hCoV-19/Wales/PHWC-48ADD6/2020\|EPI_ISL_666159\|2020 | ···················· | ···················· | ···················· | ···················· | ···················· | ···················· | |
| T | 6025 hCoV-19/Wales/PHWC-48ADE5/2020\|EPI_ISL_666160\|2020 | ···················· | ···················· | ···················· | ···················· | ···················· | ···················· | |
| T | 6026 hCoV-19/Wales/PHWC-48ADF4/2020\|EPI_ISL_666161\|2020 | ···················· | ···················· | ···················· | ···················· | ···················· | ···················· | |
| T | 6027 hCoV-19/Wales/PHWC-48AE00/2020\|EPI_ISL_666162\|2020 | ···················· | ···················· | ···················· | ···················· | ···················· | ···················· | |
| T | 6028 hCoV-19/Wales/PHWC-48AE1F/2020\|EPI_ISL_666163\|2020 | ···················· | ···················· | ···················· | ···················· | ···················· | ···················· | |
| T | 6029 hCoV-19/Wales/PHWC-48AE4C/2020\|EPI_ISL_666164\|2020 | ···················· | ···················· | ···················· | ···················· | ···················· | ···················· | |
| T | 6030 hCoV-19/Wales/PHWC-48AE6A/2020\|EPI_ISL_666165\|2020 | ···················· | ···················· | ···················· | ···················· | ···················· | ···················· | |
| T | 6031 hCoV-19/Wales/PHWC-48AE88/2020\|EPI_ISL_666166\|2020 | ···················· | ·A·················· | ···················· | ···················· | ···················· | ···················· | |
| T | 6032 hCoV-19/Wales/PHWC-48AE97/2020\|EPI_ISL_666167\|2020 | ···················· | ···················· | ···················· | ···················· | NNN················· | ···················· | 1 |
| T | 6033 hCoV-19/Wales/PHWC-48AEA6/2020\|EPI_ISL_666168\|2020 | ···················· | ···················· | ···················· | ···················· | ···················· | ···················· | |
| T | 6034 hCoV-19/Wales/PHWC-48AEB5/2020\|EPI_ISL_666169\|2020 | ···················· | ···················· | ···················· | ···················· | ···················· | ···················· | |
| T | 6035 hCoV-19/Wales/PHWC-48AEC4/2020\|EPI_ISL_666170\|2020 | ···················· | ···················· | ···················· | ···················· | ···················· | ···················· | |
| T | 6037 hCoV-19/Wales/PHWC-48AEF1/2020\|EPI_ISL_666172\|2020 | ···················· | ···················· | ···················· | ···················· | ···················· | ···················· | |
| T | 6038 hCoV-19/Wales/PHWC-48AF1C/2020\|EPI_ISL_666173\|2020 | ···················· | ···················· | ···················· | ···················· | ···················· | ···················· | |
| T | 6039 hCoV-19/Wales/PHWC-48AF2B/2020\|EPI_ISL_666174\|2020 | ···················· | ···················· | ···················· | ···················· | ···················· | ···················· | |
| T | 6040 hCoV-19/Wales/PHWC-48AF3A/2020\|EPI_ISL_666175\|2020 | ···················· | ···················· | ···················· | ···················· | ···················· | ···················· | |
| T | 6041 hCoV-19/Wales/PHWC-48AF49/2020\|EPI_ISL_666176\|2020 | ···················· | ···················· | ···················· | ···················· | ···················· | ···················· | |
| T | 6043 hCoV-19/Wales/PHWC-48AF67/2020\|EPI_ISL_666178\|2020 | ···················· | ···················· | ···················· | ···················· | ···················· | ···················· | |
| T | 6044 hCoV-19/Wales/PHWC-48AF85/2020\|EPI_ISL_666179\|2020 | ···················· | ···················· | ···················· | ···················· | ···················· | ···················· | |
| T | 6045 hCoV-19/USA/OR-OHSU-2900/2020\|EPI_ISL_667400\|2020- | ···················· | ···················· | ···················· | ···················· | ···················· | ···················· | |

| | | | | | | | |
|---|---|---|---|---|---|---|---|
| T | 6046 hCoV-19/USA/OR-OHSU-2901/2020\|EPI_ISL_667401\|2020- | ···················· | ···················· | ···················· | ···················· | ···················· | ···················· |
| T | 6047 hCoV-19/USA/OR-OHSU-2902/2020\|EPI_ISL_667402\|2020- | ···················· | ···················· | ···················· | ···················· | ···················· | ···················· |
| T | 6048 hCoV-19/USA/OR-OHSU-2903/2020\|EPI_ISL_667403\|2020- | ···················· | ···················· | ···················· | ···················· | ···················· | ···················· |
| T | 6049 hCoV-19/USA/OR-OHSU-2906/2020\|EPI_ISL_667405\|2020- | ···················· | ···················· | ···················· | ···················· | ···················· | ···················· |
| T | 6050 hCoV-19/USA/OR-OHSU-2907/2020\|EPI_ISL_667406\|2020- | ···················· | ···················· | ···················· | ···················· | ···················· | ···················· |
| T | 6052 hCoV-19/USA/WA-OHSU-2909/2020\|EPI_ISL_667408\|2020- | ···················· | ···················· | ···················· | ···················· | ···················· | ···················· |
| T | 6053 hCoV-19/USA/OR-OHSU-2910/2020\|EPI_ISL_667409\|2020- | ···················· | ···················· | ···················· | ···················· | ···················· | ···················· |
| T | 6054 hCoV-19/USA/WA-OHSU-2911/2020\|EPI_ISL_667410\|2020- | ···················· | ···················· | ···················· | ···················· | ···················· | ···················· |
| T | 6055 hCoV-19/USA/OR-OHSU-2915/2020\|EPI_ISL_667413\|2020- | ···················· | ···················· | ···················· | ···················· | ···················· | ···················· |
| T | 6056 hCoV-19/USA/OR-OHSU-2917/2020\|EPI_ISL_667414\|2020- | ···················· | ···················· | ···················· | ···················· | ···················· | ···················· |
| T | 6057 hCoV-19/USA/OR-OHSU-2918/2020\|EPI_ISL_667415\|2020- | ···················· | ·A·················· | ···················· | ···················· | ···················· | ···················· |
| T | 6058 hCoV-19/USA/CA-OHSU-2920/2020\|EPI_ISL_667417\|2020- | ···················· | ···················· | ···················· | ···················· | ···················· | ···················· |
| T | 6059 hCoV-19/USA/OR-OHSU-2923/2020\|EPI_ISL_667419\|2020- | ···················· | ···················· | ···················· | ···················· | ···················· | ···················· |
| T | 6060 hCoV-19/USA/OR-OHSU-2924/2020\|EPI_ISL_667420\|2020- | ···················· | ···················· | ···················· | ···················· | ···················· | ···················· |
| T | 6061 hCoV-19/USA/OR-OHSU-2926/2020\|EPI_ISL_667422\|2020- | ···················· | ·············A······ | ···················· | ···················· | ···················· | ···················· |
| T | 6062 hCoV-19/USA/OR-OHSU-2927/2020\|EPI_ISL_667423\|2020- | ···················· | ·A·················· | ···················· | ···················· | ···················· | ···················· |
| T | 6063 hCoV-19/USA/CA-OHSU-2929/2020\|EPI_ISL_667424\|2020- | ···················· | ·A·················· | ···················· | ···················· | ···················· | ···················· |
| T | 6064 hCoV-19/USA/OR-OHSU-2931/2020\|EPI_ISL_667426\|2020- | ···················· | ·A·················· | ···················· | ···················· | ···················· | ···················· |
| T | 6065 hCoV-19/USA/OR-OHSU-2943/2020\|EPI_ISL_667427\|2020- | ···················· | ·A·················· | ···················· | ···················· | ···················· | ···················· |
| T | 6066 hCoV-19/USA/OR-OHSU-2949/2020\|EPI_ISL_667429\|2020- | ···················· | ···················· | ···················· | ···················· | ···················· | ···················· |
| T | 6067 hCoV-19/USA/OR-OHSU-2951/2020\|EPI_ISL_667430\|2020- | ···················· | ·A·················· | ···················· | ···················· | ···················· | ···················· |
| T | 6068 hCoV-19/USA/OR-OHSU-2957/2020\|EPI_ISL_667431\|2020- | ···················· | ·A·················· | ···················· | ···················· | ···················· | ···················· |
| T | 6069 hCoV-19/USA/OR-OHSU-2971/2020\|EPI_ISL_667438\|2020- | ···················· | ·A·················· | ···················· | ···················· | ···················· | ···················· |
| T | 6070 hCoV-19/USA/OR-OHSU-2974/2020\|EPI_ISL_667439\|2020- | ···················· | ·A·················· | ···················· | ···················· | ···················· | ···················· |
| T | 6071 hCoV-19/USA/OR-OHSU-2975/2020\|EPI_ISL_667440\|2020- | ···················· | ···················· | ···················· | ···················· | ···················· | ···················· |
| T | 6072 hCoV-19/USA/OR-OHSU-2976/2020\|EPI_ISL_667441\|2020- | ···················· | ·········A·········· | ···················· | ···················· | ···················· | ···················· |
| T | 6073 hCoV-19/USA/OR-OHSU-2977/2020\|EPI_ISL_667442\|2020- | ········T··········· | ···················· | ···················· | ···················· | ···················· | ···················· |
| T | 6074 hCoV-19/USA/OR-OHSU-2980/2020\|EPI_ISL_667444\|2020- | ···················· | ···················· | ···················· | ···················· | ···················· | ···················· |
| T | 6075 hCoV-19/USA/OR-OHSU-2981/2020\|EPI_ISL_667445\|2020- | ···················· | ·A·················· | ···················· | ···················· | ···················· | ···················· |
| T | 6076 hCoV-19/USA/OR-OHSU-2982/2020\|EPI_ISL_667446\|2020- | ···················· | ···················· | ···················· | ···················· | ···················· | ···················· |
| T | 6077 hCoV-19/USA/OR-OHSU-2983/2020\|EPI_ISL_667447\|2020- | ···················· | ·A·················· | ···················· | ···················· | ···················· | ···················· |
| T | 6078 hCoV-19/USA/AZ-OHSU-2984/2020\|EPI_ISL_667448\|2020- | ···················· | ···················· | ···················· | ···················· | ···················· | ···················· |
| T | 6079 hCoV-19/USA/OR-OHSU-2986/2020\|EPI_ISL_667449\|2020- | ···················· | ·A·················· | ···················· | ···················· | ···················· | ···················· |
| T | 6080 hCoV-19/USA/OR-OHSU-2987/2020\|EPI_ISL_667450\|2020- | ···················· | ···················· | ···················· | ···················· | ···················· | ···················· |
| T | 6081 hCoV-19/USA/OR-OHSU-2990/2020\|EPI_ISL_667451\|2020- | ···················· | ·A·················· | ···················· | ···················· | ···················· | ···················· |
| T | 6082 hCoV-19/USA/OR-OHSU-2994/2020\|EPI_ISL_667452\|2020- | ···················· | ···················· | ···················· | ···················· | ···················· | ···················· |
| T | 6083 hCoV-19/USA/OR-OHSU-3043/2020\|EPI_ISL_667454\|2020- | ···················· | ·············A······ | ···················· | ···················· | ···················· | ···················· |
| T | 6084 hCoV-19/USA/OR-OHSU-3047/2020\|EPI_ISL_667457\|2020- | ···················· | ···················· | ···················· | ···················· | ···················· | ···················· |
| T | 6085 hCoV-19/USA/OR-OHSU-3048/2020\|EPI_ISL_667458\|2020- | ···················· | ···················· | ···················· | ···················· | ···················· | ···················· |
| T | 6086 hCoV-19/USA/OR-OHSU-3049/2020\|EPI_ISL_667459\|2020- | ···················· | ···················· | ···················· | ···················· | ···················· | ···················· |
| T | 6087 hCoV-19/USA/OR-OHSU-3051/2020\|EPI_ISL_667460\|2020- | ···················· | ···················· | ···················· | ···················· | ···················· | ···················· |
| T | 6088 hCoV-19/USA/OR-OHSU-3053/2020\|EPI_ISL_667462\|2020- | ···················· | ···················· | ···················· | ···················· | ···················· | ···················· |
| T | 6089 hCoV-19/USA/OR-OHSU-3058/2020\|EPI_ISL_667463\|2020- | ···················· | ·A·················· | ···················· | ···················· | ···················· | ···················· |
| T | 6090 hCoV-19/USA/OR-OHSU-3063/2020\|EPI_ISL_667465\|2020- | ···················· | ···················· | ···················· | ···················· | ···················· | ···················· |
| T | 6091 hCoV-19/USA/OR-OHSU-3065/2020\|EPI_ISL_667466\|2020- | ···················· | ···················· | ···················· | ···················· | ···················· | ···················· |
| T | 6092 hCoV-19/USA/OR-OHSU-3066/2020\|EPI_ISL_667467\|2020- | ···················· | ·········A·········· | ···················· | ···················· | ···················· | ···················· |
| T | 6093 hCoV-19/USA/OR-OHSU-3067/2020\|EPI_ISL_667468\|2020- | ···················· | ···················· | ···················· | ···················· | ···················· | ···················· |
| T | 6094 hCoV-19/USA/OR-OHSU-3071/2020\|EPI_ISL_667470\|2020- | ···················· | ···················· | ···················· | ···················· | ···················· | ···················· |
| T | 6095 hCoV-19/USA/OR-OHSU-3072/2020\|EPI_ISL_667471\|2020- | ···················· | ···················· | ···················· | ···················· | ···················· | ···················· |
| T | 6096 hCoV-19/USA/OR-OHSU-3079/2020\|EPI_ISL_667472\|2020- | ···················· | ···················· | ···················· | ···················· | ···················· | ···················· |
| T | 6097 hCoV-19/USA/OR-OHSU-3081/2020\|EPI_ISL_667473\|2020- | ···················· | ·A·················· | ···················· | ···················· | ···················· | ···················· |
| T | 6098 hCoV-19/USA/OR-OHSU-3083/2020\|EPI_ISL_667474\|2020- | ···················· | ·A·················· | ···················· | ···················· | ···················· | ···················· |
| T | 6099 hCoV-19/USA/OR-OHSU-3085/2020\|EPI_ISL_667475\|2020- | ···················· | ···················· | ···················· | ···················· | ···················· | ···················· |
| T | 6100 hCoV-19/USA/OR-OHSU-3091/2020\|EPI_ISL_667478\|2020- | ···················· | ···················· | ···················· | ···················· | ···················· | ···················· |
| T | 6101 hCoV-19/USA/OR-OHSU-3093/2020\|EPI_ISL_667479\|2020- | ···················· | ·········A·········· | ···················· | ···················· | ···················· | ···················· |
| T | 6102 hCoV-19/USA/OR-OHSU-3095/2020\|EPI_ISL_667480\|2020- | ···················· | ···················· | ···················· | ···················· | ···················· | ···················· |
| T | 6103 hCoV-19/USA/OR-OHSU-3109/2020\|EPI_ISL_667483\|2020- | ···················· | ···················· | ···················· | ···················· | ···················· | ···················· |
| T | 6104 hCoV-19/USA/OR-OHSU-3110/2020\|EPI_ISL_667484\|2020- | ···················· | ·A·················· | ···················· | ···················· | ···················· | ···················· |
| T | 6105 hCoV-19/USA/WA-OHSU-3111/2020\|EPI_ISL_667485\|2020- | ···················· | ···················· | ···················· | ···················· | ···················· | ···················· |
| T | 6106 hCoV-19/USA/OR-OHSU-3117/2020\|EPI_ISL_667488\|2020- | ···················· | ·A·················· | ···················· | ···················· | ···················· | ···················· |
| T | 6107 hCoV-19/USA/OR-OHSU-3133/2020\|EPI_ISL_667491\|2020- | ···················· | ·A·················· | ···················· | ···················· | ···················· | ···················· |
| T | 6108 hCoV-19/USA/OR-OHSU-3137/2020\|EPI_ISL_667492\|2020- | ···················· | ·A·················· | ···················· | ···················· | ···················· | ···················· |
| T | 6109 hCoV-19/USA/OR-OHSU-3139/2020\|EPI_ISL_667493\|2020- | ···················· | ·A·················· | ···················· | ···················· | ···················· | ···················· |
| T | 6110 hCoV-19/USA/OR-OHSU-3175/2020\|EPI_ISL_667498\|2020- | ···················· | ···················· | ···················· | ···················· | ···················· | ···················· |
| T | 6111 hCoV-19/Denmark/DCGC-10010/2020\|EPI_ISL_668700\|202 | ···················· | ···················· | ···················· | ···················· | ···················· | ···················· |
| T | 6112 hCoV-19/Denmark/DCGC-10023/2020\|EPI_ISL_668701\|202 | ···················· | ···················· | ···················· | ···················· | ···················· | ···················· |
| T | 6113 hCoV-19/Denmark/DCGC-10030/2020\|EPI_ISL_668702\|202 | ···················· | ···················· | ···················· | ···················· | ···················· | ···················· |
| T | 6114 hCoV-19/Denmark/DCGC-10047/2020\|EPI_ISL_668703\|202 | ···················· | ···················· | ···················· | ···················· | ···················· | ···················· |
| T | 6115 hCoV-19/Denmark/DCGC-10086/2020\|EPI_ISL_668704\|202 | ···················· | ···················· | ···················· | ···················· | ···················· | ···················· |
| T | 6116 hCoV-19/Denmark/DCGC-10090/2020\|EPI_ISL_668705\|202 | ···················· | ···················· | ···················· | ···················· | ···················· | ···················· |
| T | 6117 hCoV-19/Denmark/DCGC-10096/2020\|EPI_ISL_668706\|202 | ···················· | ···················· | ···················· | ···················· | ···················· | ···················· |
| T | 6118 hCoV-19/Denmark/DCGC-10101/2020\|EPI_ISL_668707\|202 | ···················· | ···················· | ···················· | ···················· | ···················· | ···················· |
| T | 6119 hCoV-19/Denmark/DCGC-10137/2020\|EPI_ISL_668708\|202 | ···················· | ···················· | ···················· | ···················· | ···················· | ···················· |
| T | 6120 hCoV-19/Denmark/DCGC-10157/2020\|EPI_ISL_668709\|202 | ···················· | ···················· | ···················· | ···················· | ···················· | ···················· |
| T | 6121 hCoV-19/Denmark/DCGC-10161/2020\|EPI_ISL_668710\|202 | ···················· | ···················· | ···················· | ···················· | ···················· | ···················· |
| T | 6122 hCoV-19/Denmark/DCGC-10188/2020\|EPI_ISL_668711\|202 | ···················· | ···················· | ···················· | ···················· | ···················· | ···················· |
| T | 6123 hCoV-19/Denmark/DCGC-10193/2020\|EPI_ISL_668712\|202 | ···················· | ···················· | ···················· | ···················· | ···················· | ···················· |
| T | 6124 hCoV-19/Denmark/DCGC-10199/2020\|EPI_ISL_668713\|202 | ···················· | ···················· | ···················· | ···················· | ···················· | ···················· |
| T | 6125 hCoV-19/Denmark/DCGC-9707/2020\|EPI_ISL_668714\|2020 | ···················· | ···················· | ···················· | ···················· | ···················· | ···················· |
| T | 6126 hCoV-19/Denmark/DCGC-10033/2020\|EPI_ISL_668715\|202 | ···················· | ···················· | ···················· | ···················· | ···················· | ···················· |

T 6127 hCoV-19/Denmark/DCGC-9318/2020|EPI_ISL_668718|2020 ···················· ···················· ···················· ···················· ···················· ····················
T 6128 hCoV-19/Denmark/DCGC-9321/2020|EPI_ISL_668719|2020 ···················· ···················· ···················· ···················· ···················· ····················
T 6129 hCoV-19/Denmark/DCGC-9473/2020|EPI_ISL_668720|2020 ···················· ···················· ···················· ···················· ···················· ····················
T 6130 hCoV-19/Denmark/DCGC-9557/2020|EPI_ISL_668721|2020 ···················· ···················· ···················· ···················· ···················· ····················
T 6131 hCoV-19/Denmark/DCGC-9630/2020|EPI_ISL_668722|2020 ···················· ···················· ···················· ···················· ···················· ····················
T 6132 hCoV-19/Denmark/DCGC-9717/2020|EPI_ISL_668723|2020 ···················· ···················· ···················· ···················· ···················· ····················
T 6133 hCoV-19/Denmark/DCGC-10052/2020|EPI_ISL_668724|202 ···················· ···················· ···················· ···················· ···················· ····················
T 6134 hCoV-19/Denmark/DCGC-10085/2020|EPI_ISL_668725|202 ···················· ·A·················· ···················· ···················· ···················· ··A·················
T 6135 hCoV-19/Denmark/DCGC-10095/2020|EPI_ISL_668726|202 ···················· ···················· ···················· ···················· ···················· ····················
T 6136 hCoV-19/Ecuador/UEES-7235/2020|EPI_ISL_672000|2020 ···················· ···················· ···················· ···················· ···················· ····················
T 6137 hCoV-19/Ecuador/UEES-7463/2020|EPI_ISL_672001|2020 ···················· ···················· ···················· ···················· ···················· ····················
T 6138 hCoV-19/Ecuador/UEES-32/2020|EPI_ISL_672002|2020 ···················· ···················· ···················· ···················· ···················· ····················
T 6139 hCoV-19/Ecuador/UEES-7297/2020|EPI_ISL_672004|2020 ···················· ···················· ···················· ···················· ···················· ····················
T 6140 hCoV-19/Ecuador/UEES-7493/2020|EPI_ISL_672005|2020 ···················· ···················· ···················· ···················· ···················· ····················
T 6141 hCoV-19/Ecuador/UEES-7501/2020|EPI_ISL_672008|2020 ···················· ···················· ···················· ···················· ···················· ····················
T 6142 hCoV-19/Ecuador/UEES-A001/2020|EPI_ISL_672009|2020 ···················· ···················· ···················· ···················· ···················· ····················
T 6143 hCoV-19/Ecuador/UEES-7332/2020|EPI_ISL_672011|2020 ···················· ···················· ···················· ···················· ···················· ····················
T 6144 hCoV-19/El Salvador/UES-02/2020|EPI_ISL_672012|202 ···················· ···················· ···················· ···················· ···················· ····················
T 6145 hCoV-19/USA/NY-QDX-2747/2020|EPI_ISL_661254|2020-0 ···················· ···················· ···················· ···················· ···················· ····················
T 6146 hCoV-19/New Zealand/20CV0572/2020|EPI_ISL_661258|2 ···················· ···················· ···················· ···················· ···················· ····················
T 6147 hCoV-19/New Zealand/20CV0574/2020|EPI_ISL_661259|2 ···················· ···················· ···················· ···················· ···················· ····················
T 6150 hCoV-19/New Zealand/20CV0595/2020|EPI_ISL_661265|2 ···················· ···················· ···················· ···················· ···················· ····················
T 6151 hCoV-19/Sweden/20-08949/2020|EPI_ISL_661273|2020-0 ···················· ···················· ···················· ···················· ···················· ····················
T 6152 hCoV-19/Sweden/20-09860/2020|EPI_ISL_661274|2020-1 ···················· ···················· ···················· ···················· ···················· ····················
T 6153 hCoV-19/Sweden/20-09863/2020|EPI_ISL_661275|2020-1 ···················· ···················· ···················· ···················· ···················· ····················
T 6154 hCoV-19/Sweden/20-09864/2020|EPI_ISL_661276|2020-1 ···················· ···················· ···················· ···················· ···················· ····················
T 6155 hCoV-19/Sweden/20-09865/2020|EPI_ISL_661277|2020-1 ···················· ···················· ···················· ···················· ···················· ····················
T 6156 hCoV-19/Sweden/20-22285/2020|EPI_ISL_661278|2020-0 ···················· ···················· ···················· ···················· ···················· ····················
T 6157 hCoV-19/Sweden/20-22311/2020|EPI_ISL_661279|2020-0 ···················· ···················· ···················· ···················· ···················· ····················
T 6158 hCoV-19/Sweden/20-23683/2020|EPI_ISL_661280|2020-1 ···················· ·A·················· ···················· ···················· ···················· ····················
T 6159 hCoV-19/Sweden/20-52701/2020|EPI_ISL_661281|2020-0 ···················· ···················· ···················· ···················· ···················· ····················
T 6160 hCoV-19/Sweden/20-52722/2020|EPI_ISL_661282|2020-0 ···················· ···················· ···················· ···················· ···················· ····················
T 6161 hCoV-19/Sweden/20-52819/2020|EPI_ISL_661283|2020-0 ···················· ···················· ···················· ···················· ···················· ····················
T 6162 hCoV-19/Sweden/20-52999/2020|EPI_ISL_661284|2020-0 ···················· ···················· ···················· ···················· ···················· ····················
T 6163 hCoV-19/Sweden/20-53002/2020|EPI_ISL_661285|2020-0 ···················· ···················· ···················· ···················· ···················· ····················
T 6164 hCoV-19/Sweden/20-53050/2020|EPI_ISL_661286|2020-0 ···················· ···G················ ···················· ···················· ···················· ····················
T 6165 hCoV-19/Sweden/20-53054/2020|EPI_ISL_661287|2020-0 ···················· ···················· ···················· ···················· ···················· ····················
T 6166 hCoV-19/Sweden/20-53180/2020|EPI_ISL_661288|2020-1 ···················· ···················· ···················· ···················· ···················· ····················
T 6167 hCoV-19/England/QEUH-B6B080/2020|EPI_ISL_662500|20 ···················· ·A·················· ···················· ···················· ···················· ····················
T 6168 hCoV-19/England/QEUH-B6A18E/2020|EPI_ISL_662501|20 ···················· ···················· ···················· ···················· ···················· ····················
T 6169 hCoV-19/England/ALDP-B7729A/2020|EPI_ISL_662502|20 ···················· ···················· ···················· ···················· ···················· ····················
T 6170 hCoV-19/England/QEUH-B663C8/2020|EPI_ISL_662504|20 ···················· ···················· ···················· ···················· ···················· ····················
T 6171 hCoV-19/England/QEUH-B6ADF3/2020|EPI_ISL_662505|20 ···················· ···················· ···················· ···················· ···················· ····················
T 6172 hCoV-19/England/QEUH-B6AA29/2020|EPI_ISL_662506|20 ···················· ···················· ···················· ···················· ···················· ····················
T 6173 hCoV-19/England/ALDP-B73A78/2020|EPI_ISL_662507|20 ···················· ···················· ···················· ···················· ···················· ····················
T 6174 hCoV-19/England/ALDP-B7379F/2020|EPI_ISL_662508|20 ···················· ···················· ···················· ···················· ···················· ····················
T 6175 hCoV-19/England/QEUH-B6A4D0/2020|EPI_ISL_662509|20 ···················· ···················· ···················· ···················· ···················· ····················
T 6176 hCoV-19/England/QEUH-B6A476/2020|EPI_ISL_662510|20 ···················· ···················· ···················· ···················· ···················· ····················
T 6177 hCoV-19/England/QEUH-B6B79A/2020|EPI_ISL_662512|20 ···················· ···················· ···················· ···················· ···················· ····················
T 6178 hCoV-19/England/QEUH-B6A6AD/2020|EPI_ISL_662513|20 ···················· ···················· ···················· ···················· ···················· ····················
T 6179 hCoV-19/England/ALDP-B73A3C/2020|EPI_ISL_662514|20 ···················· ···················· ···················· ···················· ···················· ····················
T 6180 hCoV-19/England/QEUH-B664B6/2020|EPI_ISL_662515|20 ···················· ·A·················· ···················· ···················· ···················· ····················
T 6181 hCoV-19/England/QEUH-B6B6E8/2020|EPI_ISL_662517|20 ···················· ···················· ···················· ···················· ···················· ····················
T 6182 hCoV-19/England/ALDP-B73744/2020|EPI_ISL_662520|20 ···················· ···················· ···················· ···················· ·····T·············· ····················
T 6183 hCoV-19/England/QEUH-B6AA47/2020|EPI_ISL_662521|20 ···················· ···················· ···················· ···················· ···················· ····················
T 6184 hCoV-19/England/QEUH-B6645C/2020|EPI_ISL_662522|20 ···················· ···················· ···················· ···················· ···················· ····················
T 6185 hCoV-19/England/QEUH-B6A485/2020|EPI_ISL_662523|20 ···················· ···················· ···················· ···················· ···················· ····················
T 6186 hCoV-19/England/QEUH-B6B0BD/2020|EPI_ISL_662525|20 ···················· ···················· ···················· ···················· ···················· ····················
T 6187 hCoV-19/England/QEUH-B6A75F/2020|EPI_ISL_662527|20 ···················· ···················· ···················· ···················· ···················· ····················
T 6188 hCoV-19/England/QEUH-B6A9FF/2020|EPI_ISL_662528|20 ···················· ···················· ···················· ···················· ···················· ····················
T 6189 hCoV-19/England/QEUH-B6AE5A/2020|EPI_ISL_662530|20 ···················· ···················· ···················· ···················· ···················· ····················
T 6190 hCoV-19/England/QEUH-B6B1E7/2020|EPI_ISL_662532|20 ···················· ···················· ···················· ···················· ···················· ····················
T 6191 hCoV-19/England/QEUH-B6A458/2020|EPI_ISL_662534|20 ···················· ···················· ···················· ···················· ···················· ····················
T 6192 hCoV-19/England/ALDP-B772B8/2020|EPI_ISL_662535|20 ···················· ···················· ···················· ···················· ···················· ····················
T 6193 hCoV-19/England/QEUH-B6A4EF/2020|EPI_ISL_662536|20 ···················· ···················· ···················· ···················· ···T················ ····················
T 6194 hCoV-19/England/QEUH-B6AA74/2020|EPI_ISL_662537|20 ···················· ···················· ···················· ···················· ·····T·············· ····················
T 6195 hCoV-19/England/QEUH-B6A6CB/2020|EPI_ISL_662538|20 ···················· ···················· ···················· ···················· ···················· ····················
T 6196 hCoV-19/England/QEUH-B67251/2020|EPI_ISL_662539|20 ···················· ···················· ···················· ···················· ···················· ····················
T 6197 hCoV-19/England/QEUH-B6B035/2020|EPI_ISL_662540|20 ···················· ···················· ···················· ···················· ···················· ····················
T 6198 hCoV-19/England/QEUH-B6AD5D/2020|EPI_ISL_662541|20 ···················· ···················· ···················· ···················· ·····T·············· ····················
T 6199 hCoV-19/England/QEUH-B6694E/2020|EPI_ISL_662542|20 ···················· ·A·················· ···················· ···················· ···················· ··A·················
T 6200 hCoV-19/England/QEUH-B6B75E/2020|EPI_ISL_662544|20 ···················· ···················· ···················· ···················· ···················· ····················
T 6201 hCoV-19/England/QEUH-B6752B/2020|EPI_ISL_662546|20 ···················· ···················· ···················· ···················· ···················· ····················
T 6202 hCoV-19/England/QEUH-B6AFFD/2020|EPI_ISL_662547|20 ···················· ···················· ···················· ···················· ···················· ····················
T 6203 hCoV-19/England/QEUH-B6AA92/2020|EPI_ISL_662548|20 ···················· ···················· ···················· ···················· ···················· ····················
T 6204 hCoV-19/England/ALDP-B735C2/2020|EPI_ISL_662549|20 ···················· ···················· ···················· ···················· ···················· ····················
T 6205 hCoV-19/England/QEUH-B6A449/2020|EPI_ISL_662550|20 ···················· ···················· ···················· ···················· ···················· ····················
T 6206 hCoV-19/England/QEUH-B66B93/2020|EPI_ISL_662551|20 ···················· ···················· ···················· ···················· ···················· ····················
T 6207 hCoV-19/England/QEUH-B6B026/2020|EPI_ISL_662552|20 ···················· ···················· ···················· ···················· ···················· ····················
T 6208 hCoV-19/England/ALDP-B73674/2020|EPI_ISL_662553|20 ···················· ···················· ···················· ···················· ···················· ····················

| | | | | | | | |
|---|---|---|---|---|---|---|---|
| T | 6209 hCoV-19/England/QEUH-B6728E/2020\|EPI_ISL_662554\|20 | ···················· | ···················· | ···················· | ···················· | ···················· | ···················· |
| T | 6210 hCoV-19/England/QEUH-B6AE4B/2020\|EPI_ISL_662555\|20 | ···················· | ···················· | ···················· | ···················· | ···················· | ···················· |
| T | 6211 hCoV-19/England/QEUH-B6A3A6/2020\|EPI_ISL_662557\|20 | ···················· | ···················· | ···················· | ···················· | ···················· | ···················· |
| T | 6212 hCoV-19/England/ALDP-B73665/2020\|EPI_ISL_662558\|20 | ···················· | ···················· | ···················· | ···················· | ···················· | ···················· |
| T | 6213 hCoV-19/England/QEUH-B6A115/2020\|EPI_ISL_662560\|20 | ···················· | ···················· | ···················· | ···················· | ···················· | ···················· |
| T | 6214 hCoV-19/England/QEUH-B66BB1/2020\|EPI_ISL_662561\|20 | ···················· | ···················· | ···················· | ···················· | ···················· | ···················· |
| T | 6215 hCoV-19/England/QEUH-B6A968/2020\|EPI_ISL_662562\|20 | ···················· | ···················· | ···················· | ···················· | ···················· | ···················· |
| T | 6216 hCoV-19/England/QEUH-B6A6E9/2020\|EPI_ISL_662563\|20 | ···················· | ·A·················· | ···················· | ···················· | ···················· | ··A················· |
| T | 6217 hCoV-19/England/ALDP-B73902/2020\|EPI_ISL_662564\|20 | ···················· | ···················· | ···················· | ···················· | ···················· | ···················· |
| T | 6218 hCoV-19/England/QEUH-B6B008/2020\|EPI_ISL_662565\|20 | ···················· | ···················· | ···················· | ···················· | ···················· | ···················· |
| T | 6219 hCoV-19/England/QEUH-B6B062/2020\|EPI_ISL_662566\|20 | ···················· | ···················· | ···················· | ···················· | ···················· | ···················· |
| T | 6220 hCoV-19/England/QEUH-B6751C/2020\|EPI_ISL_662567\|20 | ···················· | ···················· | ···················· | ···················· | ···················· | ···················· |
| T | 6221 hCoV-19/England/QEUH-B6A5BF/2020\|EPI_ISL_662568\|20 | ···················· | ···················· | ···················· | ···················· | ···················· | ···················· |
| T | 6222 hCoV-19/England/QEUH-B67567/2020\|EPI_ISL_662569\|20 | ···················· | ···················· | ···················· | ···················· | ···················· | ···················· |
| T | 6223 hCoV-19/England/QEUH-B6A4B2/2020\|EPI_ISL_662570\|20 | ···················· | ···················· | ···················· | ···················· | ···················· | ···················· |
| T | 6224 hCoV-19/England/QEUH-B66FC4/2020\|EPI_ISL_662571\|20 | ···················· | ···················· | ···················· | ···················· | ···················· | ···················· |
| T | 6225 hCoV-19/England/QEUH-B6B17E/2020\|EPI_ISL_662572\|20 | ···················· | ···················· | ···················· | ···················· | ···················· | ···················· |
| T | 6226 hCoV-19/England/QEUH-B6AE1E/2020\|EPI_ISL_662573\|20 | ···················· | ···················· | ···················· | ···················· | ···················· | ···················· |
| T | 6227 hCoV-19/England/QEUH-B6A652/2020\|EPI_ISL_662574\|20 | ···················· | ···················· | ···················· | ···················· | ···················· | ···················· |
| T | 6228 hCoV-19/England/ALDP-B7389C/2020\|EPI_ISL_662576\|20 | ···················· | ···················· | ···················· | ···················· | ···················· | ···················· |
| T | 6229 hCoV-19/England/QEUH-B66401/2020\|EPI_ISL_662577\|20 | ···················· | ···················· | ···················· | ···················· | ·····T·············· | ···················· |
| T | 6230 hCoV-19/England/ALDP-B735D1/2020\|EPI_ISL_662578\|20 | ···················· | ···················· | ···················· | ···················· | ···················· | ···················· |
| T | 6231 hCoV-19/England/ALDP-B735FF/2020\|EPI_ISL_662579\|20 | ···················· | ···················· | ···················· | ···················· | ···················· | ···················· |
| T | 6232 hCoV-19/England/QEUH-B675D0/2020\|EPI_ISL_662582\|20 | ···················· | ···················· | ···················· | ···················· | ···················· | ···················· |
| T | 6233 hCoV-19/England/ALDP-B73753/2020\|EPI_ISL_662583\|20 | ···················· | ···················· | ···················· | ···················· | ···················· | ···················· |
| T | 6234 hCoV-19/England/QEUH-B6A555/2020\|EPI_ISL_662585\|20 | ···················· | ···················· | ···················· | ···················· | ···················· | ···················· |
| T | 6235 hCoV-19/England/QEUH-B6B044/2020\|EPI_ISL_662587\|20 | ···················· | ·A·················· | ···················· | ···················· | ···················· | ··A················· |
| T | 6236 hCoV-19/England/ALDP-B7360B/2020\|EPI_ISL_662588\|20 | ···················· | ···················· | ···················· | ···················· | ···················· | ···················· |
| T | 6237 hCoV-19/England/QEUH-B6689C/2020\|EPI_ISL_662589\|20 | ···················· | ···················· | ···················· | ···················· | ···················· | ···················· |
| T | 6238 hCoV-19/England/QEUH-B6B69D/2020\|EPI_ISL_662591\|20 | ···················· | ···················· | ···················· | ···················· | ···················· | ···················· |
| T | 6239 hCoV-19/England/QEUH-B6AE78/2020\|EPI_ISL_662592\|20 | ···················· | ···················· | ···················· | ···················· | ···················· | ···················· |
| T | 6240 hCoV-19/England/ALDP-B73489/2020\|EPI_ISL_662593\|20 | ···················· | ···················· | ···················· | ···················· | ···················· | ···················· |
| T | 6241 hCoV-19/England/QEUH-B668BA/2020\|EPI_ISL_662595\|20 | ···················· | ·A·················· | ···················· | ···················· | ···················· | ··A················· |
| T | 6242 hCoV-19/England/QEUH-B6A6DA/2020\|EPI_ISL_662596\|20 | ···················· | ···················· | ···················· | ···················· | ···················· | ···················· |
| T | 6243 hCoV-19/England/ALDP-B73401/2020\|EPI_ISL_662597\|20 | ···················· | ···················· | ···················· | ···················· | ···················· | ···················· |
| T | 6244 hCoV-19/England/QEUH-B6A40D/2020\|EPI_ISL_662598\|20 | ···················· | ···················· | ···················· | ···················· | ···················· | ···················· |
| T | 6245 hCoV-19/England/QEUH-B6A230/2020\|EPI_ISL_662599\|20 | ···················· | ···················· | ···················· | ···················· | ···················· | ···················· |
| T | 6246 hCoV-19/Australia/VIC17223/2020\|EPI_ISL_663800\|202 | ···················· | ···················· | ···················· | ···················· | ···················· | ···················· |
| T | 6247 hCoV-19/Australia/VIC17224/2020\|EPI_ISL_663801\|202 | ···················· | ···················· | ···················· | ···················· | ···················· | ···················· |
| T | 6248 hCoV-19/Australia/VIC17225/2020\|EPI_ISL_663802\|202 | ···················· | ···················· | ···················· | ···················· | ···················· | ···················· |
| T | 6249 hCoV-19/Australia/VIC17226/2020\|EPI_ISL_663803\|202 | ···················· | ···················· | ···················· | ···················· | ···················· | ···················· |
| T | 6250 hCoV-19/Australia/VIC17227/2020\|EPI_ISL_663804\|202 | ···················· | ···················· | ···················· | ···················· | ···················· | ···················· |
| T | 6251 hCoV-19/Australia/VIC17228/2020\|EPI_ISL_663805\|202 | ···················· | ···················· | ···················· | ···················· | ···················· | ···················· |
| T | 6252 hCoV-19/Australia/VIC17231/2020\|EPI_ISL_663806\|202 | ···················· | ···················· | ···················· | ···················· | ···················· | ···················· |
| T | 6253 hCoV-19/Australia/VIC17233/2020\|EPI_ISL_663807\|202 | ···················· | ···················· | ···················· | ···················· | ···················· | ···················· |
| T | 6254 hCoV-19/Australia/VIC17234/2020\|EPI_ISL_663808\|202 | ···················· | ···················· | ···················· | ···················· | ···················· | ···················· |
| T | 6255 hCoV-19/Australia/VIC17235/2020\|EPI_ISL_663809\|202 | ···················· | ···················· | ···················· | ···················· | ···················· | ···················· |
| T | 6256 hCoV-19/Australia/VIC17236/2020\|EPI_ISL_663810\|202 | ···················· | ···················· | ···················· | ···················· | ···················· | ···················· |
| T | 6257 hCoV-19/Australia/VIC17237/2020\|EPI_ISL_663811\|202 | ···················· | ···················· | ···················· | ···················· | ···················· | ···················· |
| T | 6264 hCoV-19/Australia/VIC17280/2020\|EPI_ISL_663818\|202 | ···················· | ···················· | ···················· | ···················· | ···················· | ···················· |
| T | 6267 hCoV-19/Australia/VIC17291/2020\|EPI_ISL_663821\|202 | ···················· | ···················· | ···················· | ···················· | ···················· | ···················· |
| T | 6268 hCoV-19/Australia/VIC17292/2020\|EPI_ISL_663822\|202 | ···················· | ···················· | ···················· | ···················· | ···················· | ···················· |
| T | 6269 hCoV-19/Australia/VIC17293/2020\|EPI_ISL_663823\|202 | ···················· | ···················· | ···················· | ···················· | ···················· | ···················· |
| T | 6270 hCoV-19/Australia/VIC17297/2020\|EPI_ISL_663824\|202 | ···················· | ···················· | ···················· | ···················· | ···················· | ···················· |
| T | 6271 hCoV-19/Australia/VIC17299/2020\|EPI_ISL_663825\|202 | ···················· | ···················· | ···················· | ···················· | ···················· | ···················· |
| T | 6272 hCoV-19/Australia/VIC17300/2020\|EPI_ISL_663826\|202 | ···················· | ···················· | ···················· | ···················· | ···················· | ···················· |
| T | 6273 hCoV-19/Australia/VIC17302/2020\|EPI_ISL_663827\|202 | ···················· | ···················· | ···················· | ···················· | ···················· | ···················· |
| T | 6274 hCoV-19/Australia/VIC17303/2020\|EPI_ISL_663828\|202 | ···················· | ···················· | ···················· | ···················· | ···················· | ···················· |
| T | 6276 hCoV-19/Australia/VIC17305/2020\|EPI_ISL_663830\|202 | ···················· | ···················· | ···················· | ···················· | ···················· | ···················· |
| T | 6277 hCoV-19/Australia/VIC17306/2020\|EPI_ISL_663831\|202 | ···················· | ···················· | ···················· | ···················· | ···················· | ···················· |
| T | 6278 hCoV-19/Australia/VIC17307/2020\|EPI_ISL_663832\|202 | ···················· | ···················· | ···················· | ···················· | ···················· | ···················· |
| T | 6279 hCoV-19/Australia/VIC17308/2020\|EPI_ISL_663833\|202 | ···················· | ···················· | ···················· | ···················· | ···················· | ···················· |
| T | 6280 hCoV-19/Australia/VIC17311/2020\|EPI_ISL_663834\|202 | ···················· | ···················· | ···················· | ···················· | ···················· | ···················· |
| T | 6281 hCoV-19/Australia/VIC17313/2020\|EPI_ISL_663835\|202 | ···················· | ···················· | ···················· | ···················· | ···················· | ···················· |
| T | 6282 hCoV-19/Australia/VIC17314/2020\|EPI_ISL_663836\|202 | ···················· | ···················· | ···················· | ···················· | ···················· | ···················· |
| T | 6283 hCoV-19/Australia/VIC17316/2020\|EPI_ISL_663837\|202 | ···················· | ···················· | ···················· | ···················· | ···················· | ···················· |
| T | 6284 hCoV-19/Australia/VIC17319/2020\|EPI_ISL_663838\|202 | ···················· | ···················· | ···················· | ···················· | ···················· | ···················· |
| T | 6285 hCoV-19/Australia/VIC17321/2020\|EPI_ISL_663839\|202 | ···················· | ···················· | ···················· | ···················· | ···················· | ···················· |
| T | 6287 hCoV-19/Australia/VIC17326/2020\|EPI_ISL_663841\|202 | ···················· | ···················· | ···················· | ···················· | ···················· | ···················· |
| T | 6288 hCoV-19/Australia/VIC17327/2020\|EPI_ISL_663842\|202 | ···················· | ···················· | ···················· | ···················· | ···················· | ···················· |
| T | 6289 hCoV-19/Australia/VIC17330/2020\|EPI_ISL_663843\|202 | ···················· | ···················· | ···················· | ···················· | ···················· | ···················· |
| T | 6290 hCoV-19/Australia/VIC17331/2020\|EPI_ISL_663844\|202 | ···················· | ···················· | ···················· | ···················· | ···················· | ···················· |
| T | 6291 hCoV-19/Australia/VIC17332/2020\|EPI_ISL_663845\|202 | ···················· | ···················· | ···················· | ···················· | ···················· | ······W············· |
| T | 6292 hCoV-19/Australia/VIC17333/2020\|EPI_ISL_663846\|202 | ···················· | ···················· | ···················· | ···················· | ···················· | ···················· |
| T | 6293 hCoV-19/Australia/VIC17334/2020\|EPI_ISL_663847\|202 | ···················· | ···················· | ···················· | ···················· | ···················· | ···················· |
| T | 6295 hCoV-19/Australia/VIC17338/2020\|EPI_ISL_663849\|202 | ···················· | ···················· | ···················· | ···················· | ···················· | ···················· |
| T | 6296 hCoV-19/Australia/VIC17340/2020\|EPI_ISL_663850\|202 | ···················· | ···················· | ···················· | ···················· | ···················· | ···················· |
| T | 6298 hCoV-19/Australia/VIC17348/2020\|EPI_ISL_663852\|202 | ···················· | ···················· | ···················· | ···················· | ···················· | ···················· |
| T | 6299 hCoV-19/Australia/VIC17352/2020\|EPI_ISL_663853\|202 | ···················· | ···················· | ···················· | ···················· | ···················· | ···················· |
| T | 6301 hCoV-19/Australia/VIC17356/2020\|EPI_ISL_663855\|202 | ···················· | ···················· | ···················· | ···················· | ···················· | ···················· |

| | | | | | | | | |
|---|---|---|---|---|---|---|---|---|
| T | 6302 hCoV-19/Australia/VIC17359/2020\|EPI_ISL_663856\|202 | ···················· | ···················· | ···················· | ···················· | ···················· | ···················· | |
| T | 6304 hCoV-19/Australia/VIC17364/2020\|EPI_ISL_663858\|202 | ···················· | ···················· | ···················· | ···················· | ···················· | ···················· | |
| T | 6306 hCoV-19/Australia/VIC17367/2020\|EPI_ISL_663860\|202 | ···················· | ···················· | ···················· | ···················· | ···················· | ···················· | |
| T | 6308 hCoV-19/Australia/VIC17371/2020\|EPI_ISL_663862\|202 | ···················· | ···················· | ···················· | ···················· | ···················· | ···················· | |
| T | 6310 hCoV-19/Australia/VIC17375/2020\|EPI_ISL_663864\|202 | ···················· | ···················· | ···················· | ···················· | ···················· | ···················· | |
| T | 6311 hCoV-19/Australia/VIC17379/2020\|EPI_ISL_663865\|202 | ···················· | ···················· | ···················· | ···················· | ···················· | ···················· | |
| T | 6313 hCoV-19/Australia/VIC17382/2020\|EPI_ISL_663867\|202 | ···················· | ···················· | ···················· | ···················· | ···················· | ···················· | |
| T | 6314 hCoV-19/Australia/VIC17385/2020\|EPI_ISL_663868\|202 | ···················· | ···················· | ···················· | ···················· | NNN················· | ···················· | 1 |
| T | 6315 hCoV-19/Australia/VIC17386/2020\|EPI_ISL_663869\|202 | ···················· | ···················· | ···················· | ···················· | ···················· | ···················· | |
| T | 6316 hCoV-19/Australia/VIC17390/2020\|EPI_ISL_663870\|202 | ···················· | ···················· | ···················· | ···················· | ···················· | ···················· | |
| T | 6317 hCoV-19/Australia/VIC17392/2020\|EPI_ISL_663871\|202 | ···················· | ···················· | ···················· | ···················· | ···················· | ···················· | |
| T | 6318 hCoV-19/Australia/VIC17397/2020\|EPI_ISL_663872\|202 | ···················· | ···················· | ···················· | ···················· | ·····Y·············· | ···················· | |
| T | 6319 hCoV-19/Australia/VIC17399/2020\|EPI_ISL_663873\|202 | ···················· | ···················· | ···················· | ···················· | ···················· | ·······W············ | |
| T | 6320 hCoV-19/Australia/VIC17407/2020\|EPI_ISL_663874\|202 | ···················· | ···················· | ···················· | ···················· | ···················· | ···················· | |
| T | 6322 hCoV-19/Australia/VIC17411/2020\|EPI_ISL_663876\|202 | ···················· | ···················· | ···················· | ···················· | ···················· | ···················· | |
| T | 6323 hCoV-19/Australia/VIC17412/2020\|EPI_ISL_663877\|202 | ···················· | ···················· | ···················· | ···················· | ···················· | ···················· | |
| T | 6325 hCoV-19/Australia/VIC17416/2020\|EPI_ISL_663879\|202 | ···················· | ···················· | ···················· | ···················· | ···················· | ···················· | |
| T | 6326 hCoV-19/Australia/VIC17418/2020\|EPI_ISL_663880\|202 | ···················· | ···················· | ···················· | ···················· | ·Y·················· | ·············S······ | |
| T | 6330 hCoV-19/Australia/VIC17430/2020\|EPI_ISL_663884\|202 | ···················· | ···················· | ···················· | ···················· | ···················· | ···················· | |
| T | 6332 hCoV-19/Australia/VIC17434/2020\|EPI_ISL_663886\|202 | ···················· | ···················· | ···················· | ···················· | ···················· | ···················· | |
| T | 6333 hCoV-19/Australia/VIC17442/2020\|EPI_ISL_663887\|202 | ···················· | ···················· | ···················· | ···················· | ···················· | ···················· | |
| T | 6335 hCoV-19/Australia/VIC17445/2020\|EPI_ISL_663889\|202 | ···················· | ···················· | ···················· | ···················· | ···················· | ···················· | |
| T | 6337 hCoV-19/Australia/VIC17450/2020\|EPI_ISL_663891\|202 | ···················· | ···················· | ···················· | ···················· | ···················· | ···················· | |
| T | 6339 hCoV-19/Australia/VIC17454/2020\|EPI_ISL_663893\|202 | ···················· | ···················· | ···················· | ···················· | ···················· | ···················· | |
| T | 6341 hCoV-19/Australia/VIC17461/2020\|EPI_ISL_663895\|202 | ···················· | ···················· | ···················· | ···················· | ···················· | ···················· | |
| T | 6342 hCoV-19/Australia/VIC17464/2020\|EPI_ISL_663896\|202 | ···················· | ···················· | ···················· | ···················· | ···················· | ···················· | |
| T | 6343 hCoV-19/Australia/VIC17467/2020\|EPI_ISL_663897\|202 | ···················· | ···················· | ···················· | ···················· | ···················· | ···················· | |
| T | 6345 hCoV-19/Australia/VIC17475/2020\|EPI_ISL_663899\|202 | ···················· | ···················· | ···················· | ···················· | ···················· | ···················· | |
| T | 6346 hCoV-19/Scotland/EDB8981/2020\|EPI_ISL_666060\|2020- | ···················· | ···················· | ···················· | ···················· | ···················· | ···················· | |
| T | 6347 hCoV-19/Scotland/EDB9160/2020\|EPI_ISL_666061\|2020- | ···················· | ···················· | ···················· | ···················· | ···················· | ···················· | |
| T | 6349 hCoV-19/Scotland/EDB9222/2020\|EPI_ISL_666063\|2020- | ···················· | ···················· | ···················· | ···················· | ···················· | ···················· | |
| T | 6350 hCoV-19/Scotland/EDB9242/2020\|EPI_ISL_666064\|2020- | ···················· | ···················· | ···················· | ···················· | ···················· | ···················· | |
| T | 6351 hCoV-19/Scotland/EDB9261/2020\|EPI_ISL_666065\|2020- | ···················· | ···················· | ···················· | ···················· | ···················· | ···················· | |
| T | 6352 hCoV-19/Scotland/EDB9262/2020\|EPI_ISL_666066\|2020- | ···················· | ···················· | ···················· | ···················· | ···················· | ···················· | |
| T | 6353 hCoV-19/Scotland/EDB9264/2020\|EPI_ISL_666067\|2020- | ···················· | ···················· | ···················· | ···················· | ···················· | ···················· | |
| T | 6354 hCoV-19/Scotland/EDB9279/2020\|EPI_ISL_666068\|2020- | ···················· | ···················· | ···················· | ···················· | ···················· | ···················· | |
| T | 6355 hCoV-19/Scotland/EDB9280/2020\|EPI_ISL_666069\|2020- | ···················· | ···················· | ···················· | ···················· | ···················· | ···················· | |
| T | 6356 hCoV-19/Scotland/EDB9287/2020\|EPI_ISL_666070\|2020- | ···················· | ···················· | ···················· | ···················· | ···················· | ···················· | |
| T | 6357 hCoV-19/Scotland/EDB9291/2020\|EPI_ISL_666071\|2020- | ···················· | ···················· | ···················· | ···················· | ···················· | ···················· | |
| T | 6358 hCoV-19/Scotland/EDB9298/2020\|EPI_ISL_666072\|2020- | ···················· | ···················· | ···················· | ···················· | ···················· | ···················· | |
| T | 6359 hCoV-19/Scotland/EDB9299/2020\|EPI_ISL_666073\|2020- | ···················· | ···················· | ···················· | ···················· | ···················· | ···················· | |
| T | 6363 hCoV-19/Scotland/EDB9332/2020\|EPI_ISL_666077\|2020- | ···················· | ···················· | ···················· | ···················· | ···················· | ···················· | |
| T | 6364 hCoV-19/Scotland/EDB9337/2020\|EPI_ISL_666078\|2020- | ···················· | ···················· | ···················· | ···················· | ···················· | ···················· | |
| T | 6365 hCoV-19/Scotland/EDB9341/2020\|EPI_ISL_666079\|2020- | ···················· | ···················· | ···················· | ···················· | ···················· | ···················· | |
| T | 6367 hCoV-19/Scotland/EDB9353/2020\|EPI_ISL_666081\|2020- | ···················· | ···················· | ···················· | ···················· | ···················· | ···················· | |
| T | 6368 hCoV-19/Scotland/EDB9360/2020\|EPI_ISL_666082\|2020- | ···················· | ···················· | ···················· | ···················· | ···················· | ···················· | |
| T | 6369 hCoV-19/Scotland/EDB9576/2020\|EPI_ISL_666083\|2020- | ···················· | ···················· | ···················· | ···················· | ···················· | ···················· | |
| T | 6370 hCoV-19/Scotland/QEUH-A61391/2020\|EPI_ISL_666084\|2 | ···················· | ···················· | ···················· | ···················· | ···················· | ···················· | |
| T | 6371 hCoV-19/Scotland/QEUH-A613CE/2020\|EPI_ISL_666085\|2 | ···················· | ···················· | ···················· | ···················· | ···················· | ···················· | |
| T | 6372 hCoV-19/Scotland/QEUH-A61416/2020\|EPI_ISL_666086\|2 | ···················· | ···················· | ···················· | ···················· | ···················· | ···················· | |
| T | 6373 hCoV-19/Scotland/QEUH-A61C96/2020\|EPI_ISL_666087\|2 | ···················· | ···················· | ···················· | ···················· | ···················· | ···················· | |
| T | 6374 hCoV-19/Scotland/QEUH-A9F743/2020\|EPI_ISL_666088\|2 | ···················· | ···················· | ···················· | ···················· | ···················· | ···················· | |
| T | 6375 hCoV-19/Scotland/QEUH-AA05BE/2020\|EPI_ISL_666089\|2 | ···················· | ···················· | ···················· | ···················· | ···················· | ···················· | |
| T | 6376 hCoV-19/Scotland/QEUH-AA05CD/2020\|EPI_ISL_666090\|2 | ···················· | ···················· | ···················· | ···················· | ···················· | ···················· | |
| T | 6377 hCoV-19/Scotland/QEUH-AA0606/2020\|EPI_ISL_666091\|2 | ···················· | ···················· | ···················· | ···················· | ···················· | ···················· | |
| T | 6378 hCoV-19/Scotland/QEUH-AA0615/2020\|EPI_ISL_666092\|2 | ···················· | ···················· | ···················· | ···················· | ···················· | ···················· | |
| T | 6379 hCoV-19/Scotland/QEUH-AA0633/2020\|EPI_ISL_666093\|2 | ···················· | ···················· | ···················· | ···················· | ···················· | ···················· | |
| T | 6380 hCoV-19/Scotland/QEUH-AA0642/2020\|EPI_ISL_666094\|2 | ···················· | ···················· | ···················· | ···················· | ···················· | ···················· | |
| T | 6381 hCoV-19/Scotland/QEUH-AA0651/2020\|EPI_ISL_666095\|2 | ···················· | ···················· | ···················· | ···················· | ···················· | ···················· | |
| T | 6382 hCoV-19/Scotland/QEUH-AA0660/2020\|EPI_ISL_666096\|2 | ···················· | ···················· | ···················· | ···················· | ···················· | ···················· | |
| T | 6383 hCoV-19/Scotland/QEUH-AA067F/2020\|EPI_ISL_666097\|2 | ···················· | ···················· | ···················· | ···················· | ······T············· | ···················· | |
| T | 6384 hCoV-19/Scotland/QEUH-AA068E/2020\|EPI_ISL_666098\|2 | ···················· | ···················· | ···················· | ···················· | ···················· | ···················· | |
| T | 6385 hCoV-19/Scotland/QEUH-AA069D/2020\|EPI_ISL_666099\|2 | ···················· | ···················· | ···················· | ···················· | ···················· | ···················· | |
| T | 6386 hCoV-19/Scotland/QEUH-AF1141/2020\|EPI_ISL_666103\|2 | ···················· | ···················· | ···················· | ···················· | ···················· | ···················· | |
| T | 6387 hCoV-19/Scotland/QEUH-AF118D/2020\|EPI_ISL_666104\|2 | ···················· | ···················· | ···················· | ···················· | ···················· | ···················· | |
| T | 6388 hCoV-19/Scotland/QEUH-AF14EE/2020\|EPI_ISL_666106\|2 | ···················· | ···················· | ···················· | ···················· | ···················· | ···················· | |
| T | 6389 hCoV-19/USA/OR-OHSU-2889/2020\|EPI_ISL_667390\|2020- | ···················· | ·············A······ | ···················· | ···················· | ···················· | ···················· | |
| T | 6390 hCoV-19/USA/OR-OHSU-2892/2020\|EPI_ISL_667393\|2020- | ···················· | ·A·················· | ···················· | ···················· | ···················· | ···················· | |
| T | 6391 hCoV-19/USA/OR-OHSU-2895/2020\|EPI_ISL_667395\|2020- | ···················· | ·········A·········· | ···················· | ···················· | ···················· | ···················· | |
| T | 6392 hCoV-19/USA/WA-OHSU-2896/2020\|EPI_ISL_667396\|2020- | ···················· | ·A·················· | ···················· | ···················· | ···················· | ···················· | |
| T | 6393 hCoV-19/USA/WA-OHSU-2897/2020\|EPI_ISL_667397\|2020- | ···················· | ···················· | ···················· | ···················· | ···················· | ···················· | |
| T | 6394 hCoV-19/USA/OR-OHSU-2968/2020\|EPI_ISL_667435\|2020- | ···················· | ···················· | ···················· | ···················· | ···················· | ···················· | |
| T | 6395 hCoV-19/USA/OR-OHSU-2969/2020\|EPI_ISL_667436\|2020- | ···················· | ···················· | ···················· | ···················· | ···················· | ···················· | |
| T | 6396 hCoV-19/USA/OR-OHSU-2970/2020\|EPI_ISL_667437\|2020- | ···················· | ···················· | ···················· | ···················· | ···················· | ···················· | |
| T | 6397 hCoV-19/Denmark/DCGC-10420/2020\|EPI_ISL_669958\|202 | ···················· | ···················· | ···················· | ···················· | ···················· | ···················· | |
| T | 6398 hCoV-19/Denmark/DCGC-10422/2020\|EPI_ISL_669959\|202 | ···················· | ···················· | ···················· | ···················· | ···················· | ···················· | |
| T | 6399 hCoV-19/Denmark/DCGC-11484/2020\|EPI_ISL_670948\|202 | ···················· | ···················· | ···················· | ···················· | ···················· | ···················· | |
| T | 6400 hCoV-19/Denmark/DCGC-11485/2020\|EPI_ISL_670949\|202 | ···················· | ···················· | ···················· | ···················· | ···················· | ···················· | |
| T | 6401 hCoV-19/England/ALDP-B73577/2020\|EPI_ISL_662499\|20 | ···················· | ···················· | ···················· | ···················· | ···················· | ···················· | |
| T | 6402 hCoV-19/England/QEUH-AF11D8/2020\|EPI_ISL_666008\|20 | ···················· | ···················· | ···················· | ···················· | ······T············· | ···················· | |

T 6403 hCoV-19/England/QEUH-AF11E7/2020|EPI_ISL_666009|20 ···················· ···················· ···················· ···················· ···················· ····················
T 6404 hCoV-19/England/QEUH-AF1387/2020|EPI_ISL_666010|20 ···················· ···················· ···················· ···················· ···················· ····················
T 6405 hCoV-19/England/QEUH-AF13B4/2020|EPI_ISL_666011|20 ···················· ···················· ···················· ···················· ···················· ····················
T 6406 hCoV-19/England/QEUH-AF1536/2020|EPI_ISL_666013|20 ···················· ···················· ···················· ···················· ···················· ····················
T 6407 hCoV-19/England/QEUH-AF1563/2020|EPI_ISL_666014|20 ···················· ···················· ···················· ···················· ···················· ····················
T 6408 hCoV-19/England/QEUH-AF18F1/2020|EPI_ISL_666019|20 ···················· ···················· ···················· ···················· ···················· ····················
T 6409 hCoV-19/England/QEUH-AF1949/2020|EPI_ISL_666020|20 ···················· ···················· ···················· ···················· ···················· ····················
T 6410 hCoV-19/England/QEUH-AF1976/2020|EPI_ISL_666021|20 ···················· ···················· ···················· ···················· ···················· ····················
T 6411 hCoV-19/England/QEUH-B2C504/2020|EPI_ISL_666022|20 ···················· ···················· ···················· ···················· ···················· ····················
T 6412 hCoV-19/Scotland/CVR5782/2020|EPI_ISL_666023|2020- ···················· ···················· ···················· ···················· ···················· ····················
T 6413 hCoV-19/Scotland/CVR5784/2020|EPI_ISL_666024|2020- ···················· ···················· ···················· ···················· ···················· ····················
T 6414 hCoV-19/Scotland/CVR5796/2020|EPI_ISL_666025|2020- ···················· ···················· ···················· ···················· ···················· ····················
T 6415 hCoV-19/Scotland/CVR5799/2020|EPI_ISL_666026|2020- ···················· ···················· ···················· ···················· ···················· ····················
T 6416 hCoV-19/Scotland/CVR5800/2020|EPI_ISL_666027|2020- ···················· ···················· ···················· ···················· ···················· ····················
T 6417 hCoV-19/Scotland/CVR5801/2020|EPI_ISL_666028|2020- ···················· ···················· ···················· ···················· ···················· ····················
T 6418 hCoV-19/Scotland/CVR5809/2020|EPI_ISL_666029|2020- ···················· ···················· ···················· ···················· ···················· ····················
T 6419 hCoV-19/Scotland/CVR5812/2020|EPI_ISL_666030|2020- ···················· ···················· ···················· ···················· ···················· ····················
T 6420 hCoV-19/Scotland/CVR5813/2020|EPI_ISL_666031|2020- ···················· ···················· ···················· ···················· ···················· ····················
T 6421 hCoV-19/Scotland/CVR5815/2020|EPI_ISL_666032|2020- ···················· ···················· ···················· ···················· ···················· ····················
T 6422 hCoV-19/Scotland/CVR5820/2020|EPI_ISL_666033|2020- ···················· ···················· ···················· ···················· ···················· ····················
T 6423 hCoV-19/Scotland/CVR5822/2020|EPI_ISL_666034|2020- ···················· ···················· ···················· ···················· ···················· ····················
T 6424 hCoV-19/Scotland/CVR5823/2020|EPI_ISL_666035|2020- ···················· ···················· ···················· ···················· ···················· ····················
T 6425 hCoV-19/Scotland/CVR5826/2020|EPI_ISL_666036|2020- ···················· ···················· ···················· ···················· ···················· ····················
T 6426 hCoV-19/Scotland/CVR5830/2020|EPI_ISL_666037|2020- ···················· ···················· ···················· ···················· ···················· ····················
T 6427 hCoV-19/Scotland/CVR5836/2020|EPI_ISL_666038|2020- ···················· ···················· ···················· ···················· ···················· ····················
T 6428 hCoV-19/Scotland/CVR5839/2020|EPI_ISL_666039|2020- ···················· ···················· ···················· ···················· ···················· ····················
T 6429 hCoV-19/Scotland/CVR5840/2020|EPI_ISL_666040|2020- ···················· ···················· ···················· ···················· ···················· ····················
T 6430 hCoV-19/Scotland/CVR5841/2020|EPI_ISL_666041|2020- ···················· ···················· ···················· ···················· ···················· ····················
T 6431 hCoV-19/Scotland/CVR5844/2020|EPI_ISL_666042|2020- ···················· ···················· ···················· ···················· ···················· ····················
T 6432 hCoV-19/Scotland/CVR5848/2020|EPI_ISL_666043|2020- ···················· ···················· ···················· ···················· ···················· ····················
T 6433 hCoV-19/Scotland/CVR5849/2020|EPI_ISL_666044|2020- ···················· ···················· ···················· ···················· ···················· ····················
T 6434 hCoV-19/Scotland/CVR5850/2020|EPI_ISL_666045|2020- ···················· ···················· ···················· ···················· ···················· ····················
T 6435 hCoV-19/Scotland/CVR5851/2020|EPI_ISL_666046|2020- ···················· ···················· ···················· ···················· ···················· ····················
T 6436 hCoV-19/Scotland/CVR5852/2020|EPI_ISL_666047|2020- ···················· ···················· ···················· ···················· ···················· ····················
T 6437 hCoV-19/Scotland/CVR5853/2020|EPI_ISL_666048|2020- ···················· ···················· ···················· ···················· ···················· ····················
T 6438 hCoV-19/Scotland/CVR5854/2020|EPI_ISL_666049|2020- ···················· ···················· ···················· ···················· ···················· ····················
T 6439 hCoV-19/Scotland/CVR5859/2020|EPI_ISL_666050|2020- ···················· ···················· ···················· ···················· ···················· ····················
T 6440 hCoV-19/Scotland/CVR5860/2020|EPI_ISL_666051|2020- ···················· ···················· ···················· ···················· ···················· ····················
T 6441 hCoV-19/Scotland/CVR5862/2020|EPI_ISL_666052|2020- ···················· ···················· ···················· ···················· ···················· ····················
T 6442 hCoV-19/Scotland/CVR5863/2020|EPI_ISL_666053|2020- ···················· ···················· ···················· ···················· ···················· ····················
T 6443 hCoV-19/Scotland/CVR5864/2020|EPI_ISL_666054|2020- ···················· ···················· ···················· ···················· ···················· ····················
T 6444 hCoV-19/Scotland/CVR5865/2020|EPI_ISL_666055|2020- ···················· ···················· ···················· ···················· ···················· ····················
T 6445 hCoV-19/Scotland/CVR5866/2020|EPI_ISL_666056|2020- ···················· ···················· ···················· ···················· ···················· ····················
T 6446 hCoV-19/Scotland/CVR5867/2020|EPI_ISL_666057|2020- ···················· ···················· ···················· ···················· ···················· ····················
T 6447 hCoV-19/Scotland/CVR5868/2020|EPI_ISL_666058|2020- ···················· ···················· ···················· ···················· ···················· ····················
T 6448 hCoV-19/Scotland/CVR5869/2020|EPI_ISL_666059|2020- ···················· ···················· ···················· ···················· ···················· ····················
T 6449 hCoV-19/USA/OR-OHSU-2753/2020|EPI_ISL_667301|2020- ···················· ···················· ···················· ···················· ···················· ····················
T 6450 hCoV-19/USA/OR-OHSU-2765/2020|EPI_ISL_667304|2020- ···················· ···················· ···················· ···················· ···················· ····················
T 6452 hCoV-19/USA/OR-OHSU-2835/2020|EPI_ISL_667341|2020- ···················· ···················· ···················· ···················· ···················· ····················
T 6453 hCoV-19/USA/OR-OHSU-2836/2020|EPI_ISL_667342|2020- ···················· ···················· ···················· ···················· ···················· ····················
T 6454 hCoV-19/USA/OR-OHSU-2837/2020|EPI_ISL_667343|2020- ···················· ·········A·········· ···················· ···················· ···················· ····················
T 6455 hCoV-19/USA/OR-OHSU-2840/2020|EPI_ISL_667345|2020- ···················· ···················· ···················· ···················· ···················· ····················
T 6456 hCoV-19/USA/OR-OHSU-2845/2020|EPI_ISL_667350|2020- ···················· ···················· ···················· ···················· ···················· ····················
T 6457 hCoV-19/USA/OR-OHSU-2846/2020|EPI_ISL_667351|2020- ···················· ···················· ···················· ···················· ···················· ····················
T 6458 hCoV-19/USA/OR-OHSU-2849/2020|EPI_ISL_667352|2020- ···················· ···················· ···················· ···················· ···················· ····················
T 6459 hCoV-19/USA/OR-OHSU-2851/2020|EPI_ISL_667354|2020- ···················· ·A·················· ···················· ···················· ···················· ····················
T 6460 hCoV-19/USA/OR-OHSU-2852/2020|EPI_ISL_667355|2020- ···················· ···················· ···················· ···················· ···················· ····················
T 6461 hCoV-19/USA/OR-OHSU-2853/2020|EPI_ISL_667356|2020- ···················· ···················· ···················· ···················· ···················· ····················
T 6462 hCoV-19/USA/OR-OHSU-2854/2020|EPI_ISL_667357|2020- ···················· ···················· ···················· ···················· ···················· ····················
T 6463 hCoV-19/USA/OR-OHSU-2855/2020|EPI_ISL_667358|2020- ···················· ·A·················· ···················· ···················· ···················· ····················
T 6464 hCoV-19/USA/OR-OHSU-2856/2020|EPI_ISL_667359|2020- ···················· ·A·················· ···················· ···················· ···················· ····················
T 6465 hCoV-19/USA/OR-OHSU-2858/2020|EPI_ISL_667361|2020- ···················· ·A·················· ···················· ···················· ···················· ····················
T 6466 hCoV-19/USA/OR-OHSU-2860/2020|EPI_ISL_667363|2020- ···················· ···················· ···················· ···················· ···················· ····················
T 6467 hCoV-19/USA/OR-OHSU-2861/2020|EPI_ISL_667364|2020- ···················· ···················· ···················· ···················· ···················· ····················
T 6468 hCoV-19/USA/OR-OHSU-2862/2020|EPI_ISL_667365|2020- ···················· ·A·················· ···················· ···················· ···················· ····················
T 6469 hCoV-19/USA/OR-OHSU-2863/2020|EPI_ISL_667366|2020- ···················· ···················· ···················· ···················· ···················· ····················
T 6470 hCoV-19/USA/OR-OHSU-2864/2020|EPI_ISL_667367|2020- ···················· ···················· ···················· ···················· ···················· ····················
T 6471 hCoV-19/USA/OR-OHSU-2865/2020|EPI_ISL_667368|2020- ···················· ···················· ···················· ···················· ···················· ····················
T 6472 hCoV-19/USA/OR-OHSU-2866/2020|EPI_ISL_667369|2020- ···················· ·A·················· ···················· ···················· ···················· ····················
T 6473 hCoV-19/USA/OR-OHSU-2868/2020|EPI_ISL_667371|2020- ···················· ·A·················· ···················· ···················· ···················· ····················
T 6474 hCoV-19/USA/OR-OHSU-2869/2020|EPI_ISL_667372|2020- ···················· ···················· ···················· ···················· ···················· ····················
T 6475 hCoV-19/USA/OR-OHSU-2870/2020|EPI_ISL_667373|2020- ···················· ···················· ···················· ···················· ···················· ····················
T 6476 hCoV-19/USA/OR-OHSU-2871/2020|EPI_ISL_667374|2020- ···················· ···················· ···················· ···················· ···················· ····················
T 6477 hCoV-19/USA/OR-OHSU-2872/2020|EPI_ISL_667375|2020- ···················· ···················· ···················· ···················· ···················· ····················
T 6478 hCoV-19/USA/OR-OHSU-2873/2020|EPI_ISL_667376|2020- ···················· ···················· ···················· ···················· ···················· ····················
T 6479 hCoV-19/USA/OR-OHSU-2877/2020|EPI_ISL_667378|2020- ···················· ···················· ···················· ···················· ···················· ····················
T 6480 hCoV-19/USA/WA-OHSU-2879/2020|EPI_ISL_667380|2020- ···················· ·A·················· ···················· ···················· ···················· ····················
T 6481 hCoV-19/USA/WA-OHSU-2882/2020|EPI_ISL_667383|2020- ···················· ···················· ···················· ···················· ···················· ····················
T 6482 hCoV-19/USA/WA-OHSU-2883/2020|EPI_ISL_667384|2020- ···················· ···················· ···················· ···················· ···················· ····················
T 6483 hCoV-19/Denmark/DCGC-9396/2020|EPI_ISL_668601|2020 ···················· ···················· ···················· ···················· ···················· ····················

| | | | | | | | | |
|---|---|---|---|---|---|---|---|---|
| T | 6484 | hCoV-19/Denmark/DCGC-10119/2020\|EPI_ISL_668602\|202 | ····················· | ····················· | ······················ | ····················· | ····················· | ····················· |
| T | 6485 | hCoV-19/Denmark/DCGC-10129/2020\|EPI_ISL_668603\|202 | ····················· | ····················· | ······················ | ····················· | ····················· | ····················· |
| T | 6486 | hCoV-19/Denmark/DCGC-9445/2020\|EPI_ISL_668604\|2020 | ····················· | ····················· | ······················ | ····················· | ····················· | ····················· |
| T | 6487 | hCoV-19/Denmark/DCGC-9623/2020\|EPI_ISL_668605\|2020 | ····················· | ····················· | ······················ | ····················· | ····················· | ····················· |
| T | 6488 | hCoV-19/Denmark/DCGC-9229/2020\|EPI_ISL_668606\|2020 | ····················· | ····················· | ······················ | ····················· | ····················· | ····················· |
| T | 6489 | hCoV-19/Denmark/DCGC-9950/2020\|EPI_ISL_668607\|2020 | ····················· | ····················· | ······················ | ····················· | ····················· | ····················· |
| T | 6490 | hCoV-19/Denmark/DCGC-9546/2020\|EPI_ISL_668608\|2020 | ····················· | ····················· | ······················ | ····················· | ····················· | ····················· |
| T | 6491 | hCoV-19/Denmark/DCGC-9269/2020\|EPI_ISL_668609\|2020 | ····················· | ····················· | ······················ | ····················· | ····················· | ····················· |
| T | 6492 | hCoV-19/Denmark/DCGC-9308/2020\|EPI_ISL_668610\|2020 | ····················· | ····················· | ······················ | ····················· | ····················· | ····················· |
| T | 6493 | hCoV-19/Denmark/DCGC-9609/2020\|EPI_ISL_668611\|2020 | ····················· | ····················· | ······················ | ····················· | ····················· | ····················· |
| T | 6494 | hCoV-19/Denmark/DCGC-9271/2020\|EPI_ISL_668635\|2020 | ····················· | ····················· | ······················ | ····················· | ····················· | ····················· |
| T | 6495 | hCoV-19/Denmark/DCGC-9292/2020\|EPI_ISL_668636\|2020 | ····················· | ····················· | ······················ | ····················· | ····················· | ····················· |
| T | 6496 | hCoV-19/Denmark/DCGC-9310/2020\|EPI_ISL_668670\|2020 | ····················· | ····················· | ······················ | ····················· | ····················· | ····················· |
| T | 6497 | hCoV-19/Denmark/DCGC-9313/2020\|EPI_ISL_668671\|2020 | ····················· | ····················· | ······················ | ····················· | ····················· | ····················· |
| T | 6498 | hCoV-19/Denmark/DCGC-9354/2020\|EPI_ISL_668672\|2020 | ····················· | ····················· | ······················ | ····················· | ····················· | ····················· |
| T | 6499 | hCoV-19/Denmark/DCGC-9356/2020\|EPI_ISL_668673\|2020 | ····················· | ····················· | ······················ | ····················· | ····················· | ····················· |
| T | 6500 | hCoV-19/Denmark/DCGC-9392/2020\|EPI_ISL_668674\|2020 | ····················· | ····················· | ······················ | ····················· | ····················· | ····················· |
| T | 6501 | hCoV-19/Denmark/DCGC-9432/2020\|EPI_ISL_668675\|2020 | ····················· | ····················· | ······················ | ····················· | ····················· | ····················· |
| T | 6502 | hCoV-19/Denmark/DCGC-9443/2020\|EPI_ISL_668676\|2020 | ····················· | ····················· | ······················ | ····················· | ····················· | ····················· |
| T | 6503 | hCoV-19/Denmark/DCGC-9567/2020\|EPI_ISL_668680\|2020 | ····················· | ····················· | ······················ | ····················· | ····················· | ····················· |
| T | 6504 | hCoV-19/Denmark/DCGC-9575/2020\|EPI_ISL_668681\|2020 | ····················· | ····················· | ······················ | ····················· | ····················· | ····················· |
| T | 6505 | hCoV-19/Denmark/DCGC-9576/2020\|EPI_ISL_668682\|2020 | ····················· | ····················· | ······················ | ····················· | ····················· | ····················· |
| T | 6506 | hCoV-19/Denmark/DCGC-9588/2020\|EPI_ISL_668683\|2020 | ····················· | ····················· | ······················ | ····················· | ····················· | ····················· |
| T | 6507 | hCoV-19/Denmark/DCGC-9653/2020\|EPI_ISL_668684\|2020 | ····················· | ····················· | ······················ | ····················· | ····················· | ····················· |
| T | 6508 | hCoV-19/Denmark/DCGC-9654/2020\|EPI_ISL_668685\|2020 | ····················· | ····················· | ······················ | ····················· | ····················· | ····················· |
| T | 6509 | hCoV-19/Denmark/DCGC-9678/2020\|EPI_ISL_668686\|2020 | ····················· | ····················· | ······················ | ····················· | ····················· | ····················· |
| T | 6510 | hCoV-19/Denmark/DCGC-9695/2020\|EPI_ISL_668687\|2020 | ····················· | ····················· | ······················ | ····················· | ····················· | ····················· |
| T | 6511 | hCoV-19/Denmark/DCGC-9699/2020\|EPI_ISL_668688\|2020 | ····················· | ····················· | ······················ | ····················· | ····················· | ····················· |
| T | 6512 | hCoV-19/Denmark/DCGC-9702/2020\|EPI_ISL_668689\|2020 | ····················· | ····················· | ······················ | ····················· | ····················· | ····················· |
| T | 6513 | hCoV-19/Denmark/DCGC-9723/2020\|EPI_ISL_668690\|2020 | ····················· | ····················· | ······················ | ····················· | ····················· | ····················· |
| T | 6514 | hCoV-19/Denmark/DCGC-9736/2020\|EPI_ISL_668691\|2020 | ····················· | ····················· | ······················ | ····················· | ····················· | ····················· |
| T | 6515 | hCoV-19/Denmark/DCGC-9739/2020\|EPI_ISL_668692\|2020 | ····················· | ····················· | ······················ | ····················· | ····················· | ····················· |
| T | 6516 | hCoV-19/Denmark/DCGC-9829/2020\|EPI_ISL_668693\|2020 | ····················· | ····················· | ······················ | ····················· | ····················· | ····················· |
| T | 6517 | hCoV-19/Denmark/DCGC-9866/2020\|EPI_ISL_668694\|2020 | ····················· | ····················· | ······················ | ····················· | ····················· | ····················· |
| T | 6518 | hCoV-19/Denmark/DCGC-9893/2020\|EPI_ISL_668695\|2020 | ····················· | ····················· | ······················ | ····················· | ····················· | ····················· |
| T | 6519 | hCoV-19/Denmark/DCGC-9910/2020\|EPI_ISL_668696\|2020 | ····················· | ····················· | ······················ | ····················· | ····················· | ····················· |
| T | 6520 | hCoV-19/Denmark/DCGC-9911/2020\|EPI_ISL_668697\|2020 | ····················· | ····················· | ······················ | ····················· | ····················· | ····················· |
| T | 6521 | hCoV-19/Denmark/DCGC-9969/2020\|EPI_ISL_668698\|2020 | ····················· | ····················· | ······················ | ····················· | ····················· | ····················· |
| T | 6522 | hCoV-19/Denmark/DCGC-9984/2020\|EPI_ISL_668699\|2020 | ····················· | ····················· | ······················ | ····················· | ····················· | ····················· |
| T | 6523 | hCoV-19/Denmark/DCGC-10322/2020\|EPI_ISL_669900\|202 | ····················· | ·A··················· | ······················ | ····················· | ····················· | ····················· |
| T | 6524 | hCoV-19/Denmark/DCGC-10325/2020\|EPI_ISL_669901\|202 | ····················· | ····················· | ······················ | ····················· | ····················· | ····················· |
| T | 6525 | hCoV-19/Denmark/DCGC-10326/2020\|EPI_ISL_669902\|202 | ····················· | ····················· | ······················ | ····················· | ····················· | ····················· |
| T | 6526 | hCoV-19/Denmark/DCGC-10327/2020\|EPI_ISL_669903\|202 | ····················· | ····················· | ······················ | ····················· | ····················· | ····················· |
| T | 6527 | hCoV-19/Denmark/DCGC-10328/2020\|EPI_ISL_669904\|202 | ····················· | ····················· | ······················ | ····················· | ····················· | ····················· |
| T | 6528 | hCoV-19/Denmark/DCGC-10331/2020\|EPI_ISL_669905\|202 | ····················· | ····················· | ······················ | ····················· | ····················· | ····················· |
| T | 6529 | hCoV-19/Denmark/DCGC-10334/2020\|EPI_ISL_669906\|202 | ····················· | ····················· | ······················ | ····················· | ····················· | ····················· |
| T | 6530 | hCoV-19/Denmark/DCGC-10336/2020\|EPI_ISL_669907\|202 | ····················· | ····················· | ······················ | ····················· | ····················· | ····················· |
| T | 6531 | hCoV-19/Denmark/DCGC-10338/2020\|EPI_ISL_669908\|202 | ····················· | ····················· | ······················ | ····················· | ····················· | ····················· |
| T | 6532 | hCoV-19/Denmark/DCGC-10339/2020\|EPI_ISL_669909\|202 | ····················· | ····················· | ······················ | ····················· | ····················· | ····················· |
| T | 6533 | hCoV-19/Denmark/DCGC-10340/2020\|EPI_ISL_669910\|202 | ····················· | ·A··················· | ······················ | ····················· | ····················· | ··A·················· |
| T | 6534 | hCoV-19/Denmark/DCGC-10341/2020\|EPI_ISL_669911\|202 | ····················· | ····················· | ······················ | ····················· | ····················· | ····················· |
| T | 6535 | hCoV-19/Denmark/DCGC-10342/2020\|EPI_ISL_669912\|202 | ····················· | ····················· | ······················ | ····················· | ····················· | ····················· |
| T | 6536 | hCoV-19/Denmark/DCGC-10343/2020\|EPI_ISL_669913\|202 | ····················· | ····················· | ······················ | ····················· | ····················· | ····················· |
| T | 6537 | hCoV-19/Denmark/DCGC-10344/2020\|EPI_ISL_669914\|202 | ····················· | ····················· | ······················ | ····················· | ····················· | ····················· |
| T | 6538 | hCoV-19/Denmark/DCGC-10345/2020\|EPI_ISL_669915\|202 | ····················· | ····················· | ······················ | ····················· | ····················· | ····················· |
| T | 6539 | hCoV-19/Denmark/DCGC-10346/2020\|EPI_ISL_669916\|202 | ····················· | ····················· | ······················ | ····················· | ····················· | ····················· |
| T | 6540 | hCoV-19/Denmark/DCGC-10347/2020\|EPI_ISL_669917\|202 | ····················· | ····················· | ······················ | ····················· | ····················· | ····················· |
| T | 6541 | hCoV-19/Denmark/DCGC-10348/2020\|EPI_ISL_669918\|202 | ····················· | ····················· | ······················ | ····················· | ····················· | ····················· |
| T | 6542 | hCoV-19/Denmark/DCGC-10350/2020\|EPI_ISL_669919\|202 | ····················· | ····················· | ······················ | ····················· | ····················· | ····················· |
| T | 6543 | hCoV-19/Denmark/DCGC-10351/2020\|EPI_ISL_669920\|202 | ····················· | ·N··················· | ······················ | ····················· | ····················· | ····················· |
| T | 6544 | hCoV-19/Denmark/DCGC-10353/2020\|EPI_ISL_669921\|202 | ····················· | ····················· | ······················ | ····················· | ····················· | ····················· |
| T | 6545 | hCoV-19/Denmark/DCGC-10354/2020\|EPI_ISL_669922\|202 | ····················· | ····················· | ······················ | ····················· | ····················· | ····················· |
| T | 6546 | hCoV-19/Denmark/DCGC-10357/2020\|EPI_ISL_669923\|202 | ····················· | ·A··················· | ······················ | ····················· | ····················· | ····················· |
| T | 6547 | hCoV-19/Denmark/DCGC-10360/2020\|EPI_ISL_669924\|202 | ····················· | ····················· | ······················ | ····················· | ····················· | ····················· |
| T | 6548 | hCoV-19/Denmark/DCGC-10361/2020\|EPI_ISL_669925\|202 | ····················· | ····················· | ······················ | ····················· | ····················· | ····················· |
| T | 6549 | hCoV-19/Denmark/DCGC-10362/2020\|EPI_ISL_669926\|202 | ····················· | ····················· | ······················ | ····················· | ····················· | ····················· |
| T | 6550 | hCoV-19/Denmark/DCGC-10364/2020\|EPI_ISL_669927\|202 | ····················· | ····················· | ······················ | ····················· | ····················· | ····················· |
| T | 6551 | hCoV-19/Denmark/DCGC-10369/2020\|EPI_ISL_669928\|202 | ····················· | ····················· | ······················ | ····················· | ····················· | ····················· |
| T | 6552 | hCoV-19/Denmark/DCGC-10370/2020\|EPI_ISL_669929\|202 | ····················· | ····················· | ······················ | ····················· | ····················· | ····················· |
| T | 6553 | hCoV-19/Denmark/DCGC-10375/2020\|EPI_ISL_669930\|202 | ····················· | ····················· | ······················ | ····················· | ····················· | ····················· |
| T | 6554 | hCoV-19/Denmark/DCGC-10376/2020\|EPI_ISL_669931\|202 | ····················· | ····················· | ······················ | ····················· | ····················· | ····················· |
| T | 6555 | hCoV-19/Denmark/DCGC-10378/2020\|EPI_ISL_669932\|202 | ····················· | ····················· | ······················ | ····················· | ····················· | ····················· |
| T | 6556 | hCoV-19/Denmark/DCGC-10382/2020\|EPI_ISL_669933\|202 | ····················· | ····················· | ······················ | ····················· | ····················· | ····················· |
| T | 6557 | hCoV-19/Denmark/DCGC-10384/2020\|EPI_ISL_669934\|202 | ····················· | ····················· | ······················ | ····················· | ····················· | ····················· |
| T | 6558 | hCoV-19/Denmark/DCGC-10385/2020\|EPI_ISL_669935\|202 | ····················· | ····················· | ······················ | ····················· | ····················· | ····················· |
| T | 6559 | hCoV-19/Denmark/DCGC-10386/2020\|EPI_ISL_669936\|202 | ····················· | ····················· | ······················ | ····················· | ····················· | ····················· |
| T | 6560 | hCoV-19/Denmark/DCGC-10389/2020\|EPI_ISL_669937\|202 | ····················· | ····················· | ······················ | ····················· | ····················· | ····················· |
| T | 6561 | hCoV-19/Denmark/DCGC-10391/2020\|EPI_ISL_669938\|202 | ····················· | ····················· | ······················ | ····················· | ····················· | ····················· |
| T | 6562 | hCoV-19/Denmark/DCGC-10393/2020\|EPI_ISL_669939\|202 | ····················· | ····················· | ······················ | ····················· | ····················· | ····················· |
| T | 6563 | hCoV-19/Denmark/DCGC-10394/2020\|EPI_ISL_669940\|202 | ····················· | ····················· | ······················ | ····················· | ·····T··············· | ····················· |

| | | | | | | | |
|---|---|---|---|---|---|---|---|
| T | 6564 hCoV-19/Denmark/DCGC-10396/2020\|EPI_ISL_669941\|202 | ···················· | ···················· | ····················· | ···················· | ···················· | ···················· |
| T | 6565 hCoV-19/Denmark/DCGC-10397/2020\|EPI_ISL_669942\|202 | ···················· | ···················· | ····················· | ···················· | ···················· | ···················· |
| T | 6566 hCoV-19/Denmark/DCGC-10398/2020\|EPI_ISL_669943\|202 | ···················· | ···················· | ····················· | ···················· | ···················· | ···················· |
| T | 6567 hCoV-19/Denmark/DCGC-10399/2020\|EPI_ISL_669944\|202 | ···················· | ···················· | ····················· | ···················· | ···················· | ···················· |
| T | 6568 hCoV-19/Denmark/DCGC-10400/2020\|EPI_ISL_669945\|202 | ···················· | ·A·················· | ····················· | ···················· | ···················· | ···················· |
| T | 6569 hCoV-19/Denmark/DCGC-10401/2020\|EPI_ISL_669946\|202 | ···················· | ···················· | ····················· | ···················· | ···················· | ···················· |
| T | 6570 hCoV-19/Denmark/DCGC-10402/2020\|EPI_ISL_669947\|202 | ···················· | ···················· | ····················· | ···················· | ···················· | ···················· |
| T | 6571 hCoV-19/Denmark/DCGC-10403/2020\|EPI_ISL_669948\|202 | ···················· | ···················· | ····················· | ···················· | ···················· | ···················· |
| T | 6572 hCoV-19/Denmark/DCGC-10407/2020\|EPI_ISL_669949\|202 | ···················· | ···················· | ····················· | ···················· | ···················· | ···················· |
| T | 6573 hCoV-19/Denmark/DCGC-10408/2020\|EPI_ISL_669950\|202 | ···················· | ···················· | ····················· | ···················· | ···················· | ···················· |
| T | 6574 hCoV-19/Denmark/DCGC-10410/2020\|EPI_ISL_669951\|202 | ···················· | ···················· | ····················· | ···················· | ···················· | ···················· |
| T | 6575 hCoV-19/Denmark/DCGC-10411/2020\|EPI_ISL_669952\|202 | ···················· | ···················· | ····················· | ···················· | ···················· | ···················· |
| T | 6576 hCoV-19/Denmark/DCGC-10412/2020\|EPI_ISL_669953\|202 | ···················· | ···················· | ····················· | ···················· | ···················· | ···················· |
| T | 6577 hCoV-19/Denmark/DCGC-10413/2020\|EPI_ISL_669954\|202 | ···················· | ···················· | ····················· | ···················· | ···················· | ···················· |
| T | 6578 hCoV-19/Denmark/DCGC-10415/2020\|EPI_ISL_669955\|202 | ···················· | ···················· | ····················· | ···················· | ···················· | ···················· |
| T | 6579 hCoV-19/Denmark/DCGC-10416/2020\|EPI_ISL_669956\|202 | ···················· | ···················· | ····················· | ···················· | ···················· | ···················· |
| T | 6580 hCoV-19/Denmark/DCGC-10417/2020\|EPI_ISL_669957\|202 | ···················· | ·A·················· | ····················· | ···················· | ···················· | ···················· |
| T | 6581 hCoV-19/Denmark/DCGC-10423/2020\|EPI_ISL_669960\|202 | ···················· | ···················· | ····················· | ···················· | ···················· | ···················· |
| T | 6582 hCoV-19/Denmark/DCGC-10426/2020\|EPI_ISL_669961\|202 | ···················· | ···················· | ····················· | ···················· | ···················· | ···················· |
| T | 6583 hCoV-19/Denmark/DCGC-10427/2020\|EPI_ISL_669962\|202 | ···················· | ·A·················· | ····················· | ···················· | ···················· | ···················· |
| T | 6584 hCoV-19/Denmark/DCGC-10428/2020\|EPI_ISL_669963\|202 | ···················· | ···················· | ····················· | ···················· | ···················· | ···················· |
| T | 6585 hCoV-19/Denmark/DCGC-10429/2020\|EPI_ISL_669964\|202 | ···················· | ···················· | ····················· | ···················· | ···················· | ···················· |
| T | 6586 hCoV-19/Denmark/DCGC-10431/2020\|EPI_ISL_669965\|202 | ···················· | ···················· | ····················· | ···················· | ···················· | ···················· |
| T | 6587 hCoV-19/Denmark/DCGC-10432/2020\|EPI_ISL_669966\|202 | ···················· | ···················· | ····················· | ···················· | ···················· | ···················· |
| T | 6588 hCoV-19/Denmark/DCGC-10434/2020\|EPI_ISL_669967\|202 | ···················· | ···················· | ····················· | ···················· | ···················· | ···················· |
| T | 6589 hCoV-19/Denmark/DCGC-11411/2020\|EPI_ISL_670900\|202 | ···················· | ···················· | ····················· | ···················· | ···················· | ···················· |
| T | 6590 hCoV-19/Denmark/DCGC-11412/2020\|EPI_ISL_670901\|202 | ···················· | ···················· | ····················· | ···················· | ···················· | ···················· |
| T | 6591 hCoV-19/Denmark/DCGC-11413/2020\|EPI_ISL_670902\|202 | ···················· | ···················· | ····················· | ···················· | ······T············· | ···················· |
| T | 6592 hCoV-19/Denmark/DCGC-11414/2020\|EPI_ISL_670903\|202 | ···················· | ···················· | ····················· | ···················· | ···················· | ···················· |
| T | 6593 hCoV-19/Denmark/DCGC-11415/2020\|EPI_ISL_670904\|202 | ···················· | ·A·················· | ····················· | ···················· | ···················· | ···················· |
| T | 6594 hCoV-19/Denmark/DCGC-11416/2020\|EPI_ISL_670905\|202 | ···················· | ···················· | ····················· | ···················· | ···················· | ···················· |
| T | 6595 hCoV-19/Denmark/DCGC-11417/2020\|EPI_ISL_670906\|202 | ···················· | ···················· | ····················· | ···················· | ···················· | ···················· |
| T | 6596 hCoV-19/Denmark/DCGC-11418/2020\|EPI_ISL_670907\|202 | ···················· | ·A·················· | ····················· | ···················· | ···················· | ···A················ |
| T | 6597 hCoV-19/Denmark/DCGC-11419/2020\|EPI_ISL_670908\|202 | ···················· | ···················· | ····················· | ···················· | ···················· | ···················· |
| T | 6598 hCoV-19/Denmark/DCGC-11420/2020\|EPI_ISL_670909\|202 | ···················· | ···················· | ····················· | ···················· | ···················· | ···················· |
| T | 6599 hCoV-19/Denmark/DCGC-11422/2020\|EPI_ISL_670910\|202 | ···················· | ···················· | ····················· | ···················· | ···················· | ···················· |
| T | 6600 hCoV-19/Denmark/DCGC-11423/2020\|EPI_ISL_670911\|202 | ···················· | ···················· | ····················· | ···················· | ···················· | ···················· |
| T | 6601 hCoV-19/Denmark/DCGC-11425/2020\|EPI_ISL_670912\|202 | ···················· | ···················· | ····················· | ···················· | ···················· | ···················· |
| T | 6602 hCoV-19/Denmark/DCGC-11427/2020\|EPI_ISL_670913\|202 | ···················· | ···················· | ····················· | ···················· | ···················· | ···················· |
| T | 6603 hCoV-19/Denmark/DCGC-11428/2020\|EPI_ISL_670914\|202 | ···················· | ···················· | ····················· | ···················· | ···················· | ···················· |
| T | 6604 hCoV-19/Denmark/DCGC-11429/2020\|EPI_ISL_670915\|202 | ···················· | ···················· | ····················· | ···················· | ···················· | ···················· |
| T | 6605 hCoV-19/Denmark/DCGC-11430/2020\|EPI_ISL_670916\|202 | ···················· | ···················· | ····················· | ···················· | ···················· | ···················· |
| T | 6606 hCoV-19/Denmark/DCGC-11431/2020\|EPI_ISL_670917\|202 | ···················· | ···················· | ····················· | ···················· | ···················· | ···················· |
| T | 6607 hCoV-19/Denmark/DCGC-11432/2020\|EPI_ISL_670918\|202 | ···················· | ···················· | ····················· | ···················· | ···················· | ···················· |
| T | 6608 hCoV-19/Denmark/DCGC-11434/2020\|EPI_ISL_670919\|202 | ···················· | ···················· | ····················· | ···················· | ···················· | ···················· |
| T | 6609 hCoV-19/Denmark/DCGC-11435/2020\|EPI_ISL_670920\|202 | ···················· | ···················· | ····················· | ···················· | ···················· | ···················· |
| T | 6610 hCoV-19/Denmark/DCGC-11437/2020\|EPI_ISL_670921\|202 | ···················· | ···················· | ····················· | ···················· | ···················· | ···················· |
| T | 6611 hCoV-19/Denmark/DCGC-11441/2020\|EPI_ISL_670922\|202 | ···················· | ····A··············· | ····················· | ···················· | ···················· | ···················· |
| T | 6612 hCoV-19/Denmark/DCGC-11442/2020\|EPI_ISL_670923\|202 | ···················· | ·A·················· | ····················· | ···················· | ···················· | ···················· |
| T | 6613 hCoV-19/Denmark/DCGC-11444/2020\|EPI_ISL_670924\|202 | ···················· | ···················· | ····················· | ···················· | ···················· | ···················· |
| T | 6614 hCoV-19/Denmark/DCGC-11445/2020\|EPI_ISL_670925\|202 | ···················· | ···················· | ····················· | ···················· | ···················· | ···················· |
| T | 6615 hCoV-19/Denmark/DCGC-11446/2020\|EPI_ISL_670926\|202 | ···················· | ···················· | ····················· | ···················· | ···················· | ···················· |
| T | 6616 hCoV-19/Denmark/DCGC-11447/2020\|EPI_ISL_670927\|202 | ···················· | ···················· | ····················· | ···················· | ···················· | ···················· |
| T | 6617 hCoV-19/Denmark/DCGC-11449/2020\|EPI_ISL_670928\|202 | ···················· | ···················· | ····················· | ···················· | ···················· | ···················· |
| T | 6618 hCoV-19/Denmark/DCGC-11452/2020\|EPI_ISL_670929\|202 | ···················· | ···················· | ····················· | ···················· | ···················· | ···················· |
| T | 6619 hCoV-19/Denmark/DCGC-11453/2020\|EPI_ISL_670930\|202 | ···················· | ···················· | ····················· | ···················· | ···················· | ···················· |
| T | 6620 hCoV-19/Denmark/DCGC-11456/2020\|EPI_ISL_670931\|202 | ···················· | ···················· | ····················· | ···················· | ···················· | ···················· |
| T | 6621 hCoV-19/Denmark/DCGC-11461/2020\|EPI_ISL_670932\|202 | ···················· | ·A·················· | ····················· | ···················· | ···················· | ···················· |
| T | 6622 hCoV-19/Denmark/DCGC-11466/2020\|EPI_ISL_670933\|202 | ···················· | ···················· | ····················· | ···················· | ···················· | ···················· |
| T | 6623 hCoV-19/Denmark/DCGC-11468/2020\|EPI_ISL_670934\|202 | ···················· | ···················· | ····················· | ···················· | ···················· | ···················· |
| T | 6624 hCoV-19/Denmark/DCGC-11469/2020\|EPI_ISL_670935\|202 | ···················· | ···················· | ····················· | ···················· | ···················· | ···················· |
| T | 6625 hCoV-19/Denmark/DCGC-11470/2020\|EPI_ISL_670936\|202 | ···················· | ···················· | ····················· | ···················· | ···················· | ···················· |
| T | 6626 hCoV-19/Denmark/DCGC-11471/2020\|EPI_ISL_670937\|202 | ···················· | ···················· | ····················· | ···················· | ···················· | ···················· |
| T | 6627 hCoV-19/Denmark/DCGC-11472/2020\|EPI_ISL_670938\|202 | ···················· | ···················· | ····················· | ···················· | ···················· | ···················· |
| T | 6628 hCoV-19/Denmark/DCGC-11474/2020\|EPI_ISL_670939\|202 | ···················· | ···················· | ····················· | ···················· | ···················· | ···················· |
| T | 6629 hCoV-19/Denmark/DCGC-11475/2020\|EPI_ISL_670940\|202 | ···················· | ···················· | ····················· | ···················· | ···················· | ···················· |
| T | 6630 hCoV-19/Denmark/DCGC-11476/2020\|EPI_ISL_670941\|202 | ···················· | ·A·················· | ····················· | ···················· | ···················· | ···A················ |
| T | 6631 hCoV-19/Denmark/DCGC-11477/2020\|EPI_ISL_670942\|202 | ···················· | ···················· | ····················· | ···················· | ···················· | ···················· |
| T | 6632 hCoV-19/Denmark/DCGC-11478/2020\|EPI_ISL_670943\|202 | ···················· | ···················· | ····················· | ···················· | ···················· | ···················· |
| T | 6633 hCoV-19/Denmark/DCGC-11479/2020\|EPI_ISL_670944\|202 | ···················· | ···················· | ····················· | ···················· | ···················· | ···················· |
| T | 6634 hCoV-19/Denmark/DCGC-11480/2020\|EPI_ISL_670945\|202 | ···················· | ···················· | ····················· | ···················· | ······T············· | ···················· |
| T | 6635 hCoV-19/Denmark/DCGC-11482/2020\|EPI_ISL_670946\|202 | ···················· | ···················· | ····················· | ···················· | ···················· | ···················· |
| T | 6636 hCoV-19/Denmark/DCGC-11483/2020\|EPI_ISL_670947\|202 | ···················· | ·········A·········· | ····················· | ···················· | ···················· | ···················· |
| T | 6637 hCoV-19/Denmark/DCGC-11535/2020\|EPI_ISL_670980\|202 | ···················· | ···················· | ····················· | ···················· | ···················· | ···················· |
| T | 6638 hCoV-19/Denmark/DCGC-11536/2020\|EPI_ISL_670981\|202 | ···················· | ···················· | ····················· | ···················· | ···················· | ···················· |
| T | 6639 hCoV-19/Denmark/DCGC-11537/2020\|EPI_ISL_670982\|202 | ···················· | ···················· | ····················· | ···················· | ···················· | ···················· |
| T | 6640 hCoV-19/Denmark/DCGC-11538/2020\|EPI_ISL_670983\|202 | ···················· | ···················· | ····················· | ···················· | ···················· | ···················· |
| T | 6641 hCoV-19/Denmark/DCGC-11540/2020\|EPI_ISL_670984\|202 | ···················· | ···················· | ····················· | ···················· | ···················· | ···················· |
| T | 6642 hCoV-19/Denmark/DCGC-11542/2020\|EPI_ISL_670985\|202 | ···················· | ···················· | ····················· | ···················· | ···················· | ···················· |
| T | 6643 hCoV-19/Denmark/DCGC-11544/2020\|EPI_ISL_670986\|202 | ···················· | ···················· | ····················· | ···················· | ···················· | ···················· |

| | | | | | | | |
|---|---|---|---|---|---|---|---|
| T | 6644 hCoV-19/Denmark/DCGC-11545/2020\|EPI_ISL_670987\|202 | ···················· | ···················· | ···················· | ···················· | ···················· | ···················· |
| T | 6645 hCoV-19/Denmark/DCGC-11547/2020\|EPI_ISL_670988\|202 | ···················· | ···················· | ···················· | ···················· | ···················· | ···················· |
| T | 6646 hCoV-19/Denmark/DCGC-11552/2020\|EPI_ISL_670989\|202 | ···················· | ···················· | ···················· | ···················· | ···················· | ···················· |
| T | 6647 hCoV-19/Denmark/DCGC-11553/2020\|EPI_ISL_670990\|202 | ···················· | ···················· | ···················· | ···················· | ···················· | ···················· |
| T | 6648 hCoV-19/Denmark/DCGC-11555/2020\|EPI_ISL_670991\|202 | ···················· | ···················· | ···················· | ···················· | ···················· | ···················· |
| T | 6649 hCoV-19/Denmark/DCGC-11556/2020\|EPI_ISL_670992\|202 | ···················· | ···················· | ···················· | ···················· | ···················· | ···················· |
| T | 6650 hCoV-19/Denmark/DCGC-11558/2020\|EPI_ISL_670993\|202 | ···················· | ···················· | ···················· | ···················· | ···················· | ···················· |
| T | 6651 hCoV-19/Denmark/DCGC-11559/2020\|EPI_ISL_670994\|202 | ···················· | ·A·················· | ···················· | ···················· | ···················· | ···················· |
| T | 6652 hCoV-19/Denmark/DCGC-11560/2020\|EPI_ISL_670995\|202 | ···················· | ···················· | ···················· | ···················· | ···················· | ···················· |
| T | 6653 hCoV-19/Denmark/DCGC-11561/2020\|EPI_ISL_670996\|202 | ···················· | ···················· | ···················· | ···················· | ···················· | ···················· |
| T | 6654 hCoV-19/Denmark/DCGC-11565/2020\|EPI_ISL_670997\|202 | ···················· | ···················· | ···················· | ···················· | ···················· | ···················· |
| T | 6655 hCoV-19/Denmark/DCGC-11566/2020\|EPI_ISL_670998\|202 | ···················· | ···················· | ···················· | ···················· | ···················· | ···················· |
| T | 6656 hCoV-19/Denmark/DCGC-11569/2020\|EPI_ISL_670999\|202 | ···················· | ···················· | ···················· | ···················· | ···················· | ···················· |
| T | 6657 hCoV-19/England/QEUH-B6B6AC/2020\|EPI_ISL_662401\|20 | ···················· | ···················· | ···················· | ···················· | ···················· | ···················· |
| T | 6658 hCoV-19/England/ALDP-B73683/2020\|EPI_ISL_662405\|20 | ···················· | ···················· | ···················· | ···················· | ···················· | ···················· |
| T | 6659 hCoV-19/England/QEUH-B66902/2020\|EPI_ISL_662406\|20 | ···················· | ···················· | ···················· | ···················· | ···················· | ···················· |
| T | 6660 hCoV-19/England/QEUH-B6B132/2020\|EPI_ISL_662407\|20 | ···················· | ···················· | ···················· | ···················· | ···················· | ···················· |
| T | 6661 hCoV-19/England/QEUH-B672BB/2020\|EPI_ISL_662408\|20 | ···················· | ···················· | ···················· | ···················· | ···················· | ···················· |
| T | 6662 hCoV-19/England/QEUH-B6B74F/2020\|EPI_ISL_662409\|20 | ···················· | ···················· | ···················· | ···················· | ···················· | ···················· |
| T | 6663 hCoV-19/England/ALDP-B73A2D/2020\|EPI_ISL_662433\|20 | ···················· | ···················· | ···················· | ···················· | ···················· | ···················· |
| T | 6664 hCoV-19/England/QEUH-B6A573/2020\|EPI_ISL_662434\|20 | ···················· | ···················· | ···················· | ···················· | ···················· | ···················· |
| T | 6665 hCoV-19/England/ALDP-B73A1E/2020\|EPI_ISL_662446\|20 | ···················· | ···················· | ···················· | ···················· | ···················· | ···················· |
| T | 6666 hCoV-19/England/QEUH-B6A36A/2020\|EPI_ISL_662448\|20 | ···················· | ···················· | ···················· | ···················· | ···················· | ···················· |
| T | 6667 hCoV-19/England/QEUH-B6B6D9/2020\|EPI_ISL_662449\|20 | ···················· | ···················· | ···················· | ···················· | ···················· | ···················· |
| T | 6668 hCoV-19/England/QEUH-B6A9A4/2020\|EPI_ISL_662450\|20 | ···················· | ···················· | ···················· | ···················· | ···················· | ···················· |
| T | 6669 hCoV-19/England/QEUH-B6A77D/2020\|EPI_ISL_662451\|20 | ···················· | ···················· | ···················· | ···················· | ···················· | ···················· |
| T | 6670 hCoV-19/England/ALDP-B735A4/2020\|EPI_ISL_662452\|20 | ···················· | ···················· | ···················· | ···················· | ···················· | ···················· |
| T | 6671 hCoV-19/England/QEUH-B66B75/2020\|EPI_ISL_662453\|20 | ···················· | ···················· | ···················· | ···················· | ···················· | ···················· |
| T | 6672 hCoV-19/England/QEUH-B675B2/2020\|EPI_ISL_662454\|20 | ···················· | ···················· | ···················· | ···················· | ···················· | ···················· |
| T | 6673 hCoV-19/England/QEUH-B6736D/2020\|EPI_ISL_662455\|20 | ···················· | ···················· | ···················· | ···················· | ···················· | ···················· |
| T | 6674 hCoV-19/England/QEUH-B6A467/2020\|EPI_ISL_662456\|20 | ···················· | ···················· | ···················· | ···················· | ···················· | ···················· |
| T | 6675 hCoV-19/England/ALDP-B772D6/2020\|EPI_ISL_662457\|20 | ···················· | ···················· | ···················· | ···················· | ···················· | ···················· |
| T | 6676 hCoV-19/England/QEUH-B6B0EA/2020\|EPI_ISL_662458\|20 | ···················· | ···················· | ···················· | ···················· | ···················· | ···················· |
| T | 6677 hCoV-19/England/QEUH-B6654A/2020\|EPI_ISL_662459\|20 | ···················· | ···················· | ···················· | ···················· | ···················· | ···················· |
| T | 6678 hCoV-19/England/QEUH-B6A704/2020\|EPI_ISL_662461\|20 | ···················· | ···················· | ···················· | ···················· | ···················· | ···················· |
| T | 6679 hCoV-19/England/QEUH-B66B48/2020\|EPI_ISL_662465\|20 | ···················· | ···················· | ···················· | ···················· | ···················· | ···················· |
| T | 6680 hCoV-19/England/QEUH-B6A160/2020\|EPI_ISL_662466\|20 | ···················· | ···················· | ···················· | ···················· | ···················· | ···················· |
| T | 6681 hCoV-19/England/QEUH-B6A546/2020\|EPI_ISL_662467\|20 | ···················· | ···················· | ···················· | ···················· | ···················· | ···················· |
| T | 6682 hCoV-19/England/ALDP-B734E3/2020\|EPI_ISL_662468\|20 | ···················· | ···················· | ···················· | ···················· | ···················· | ···················· |
| T | 6683 hCoV-19/England/QEUH-B6A670/2020\|EPI_ISL_662469\|20 | ···················· | ···················· | ···················· | ···················· | ···················· | ···················· |
| T | 6684 hCoV-19/England/QEUH-B6A713/2020\|EPI_ISL_662471\|20 | ···················· | ···················· | ···················· | ···················· | ···················· | ···················· |
| T | 6685 hCoV-19/England/ALDP-B739F3/2020\|EPI_ISL_662472\|20 | ···················· | ·A·················· | ···················· | ···················· | ···················· | ···················· |
| T | 6686 hCoV-19/England/QEUH-B6A7AA/2020\|EPI_ISL_662473\|20 | ···················· | ···················· | ···················· | ···················· | ···················· | ···················· |
| T | 6687 hCoV-19/England/ALDP-B7345C/2020\|EPI_ISL_662474\|20 | ···················· | ···················· | ···················· | ···················· | ···················· | ···················· |
| T | 6688 hCoV-19/England/QEUH-B6A7D7/2020\|EPI_ISL_662475\|20 | ···················· | ···················· | ···················· | ···················· | ···················· | ···················· |
| T | 6689 hCoV-19/England/QEUH-B66B0C/2020\|EPI_ISL_662476\|20 | ···················· | ···················· | ···················· | ···················· | ···················· | ···················· |
| T | 6690 hCoV-19/England/ALDP-B73586/2020\|EPI_ISL_662477\|20 | ···················· | ···················· | ···················· | ···················· | ···················· | ···················· |
| T | 6691 hCoV-19/England/ALDP-B73920/2020\|EPI_ISL_662478\|20 | ···················· | ···················· | ···················· | ···················· | ···················· | ···················· |
| T | 6692 hCoV-19/England/ALDP-B73A87/2020\|EPI_ISL_662480\|20 | ···················· | ···················· | ···················· | ···················· | ···················· | ···················· |
| T | 6693 hCoV-19/England/QEUH-B6B09F/2020\|EPI_ISL_662481\|20 | ···················· | ···················· | ···················· | ···················· | ···················· | ···················· |
| T | 6694 hCoV-19/England/QEUH-B6A3F1/2020\|EPI_ISL_662482\|20 | ···················· | ···················· | ···················· | ···················· | ···················· | ···················· |
| T | 6695 hCoV-19/England/QEUH-B672CA/2020\|EPI_ISL_662484\|20 | ···················· | ···················· | ···················· | ···················· | ···················· | ···················· |
| T | 6696 hCoV-19/England/ALDP-B73629/2020\|EPI_ISL_662486\|20 | ···················· | ···················· | ···················· | ···················· | ···················· | ···················· |
| T | 6697 hCoV-19/England/ALDP-B7727C/2020\|EPI_ISL_662487\|20 | ···················· | ···················· | ···················· | ···················· | ···················· | ···················· |
| T | 6698 hCoV-19/Scotland/ALDP-B73850/2020\|EPI_ISL_662488\|2 | ···················· | ···················· | ···················· | ···················· | ···················· | ···················· |
| T | 6699 hCoV-19/England/QEUH-B6B1D8/2020\|EPI_ISL_662490\|20 | ···················· | ·A·················· | ···················· | ···················· | ···················· | ··A················· |
| T | 6700 hCoV-19/England/ALDP-B73568/2020\|EPI_ISL_662492\|20 | ···················· | ···················· | ···················· | ···················· | ···················· | ···················· |
| T | 6701 hCoV-19/England/QEUH-B6B76D/2020\|EPI_ISL_662494\|20 | ···················· | ···················· | ···················· | ···················· | ···················· | ···················· |
| T | 6702 hCoV-19/England/QEUH-B6A43A/2020\|EPI_ISL_662495\|20 | ···················· | ···················· | ···················· | ···················· | ···················· | ···················· |
| T | 6703 hCoV-19/England/ALDP-B735E0/2020\|EPI_ISL_662496\|20 | ···················· | ···················· | ···················· | ···················· | ···················· | ···················· |
| T | 6704 hCoV-19/England/QEUH-B6A34C/2020\|EPI_ISL_662497\|20 | ···················· | ···················· | ···················· | ···················· | ···················· | ···················· |
| T | 6705 hCoV-19/England/QEUH-B6A5A0/2020\|EPI_ISL_662498\|20 | ···················· | ···················· | ···················· | ···················· | ···················· | ···················· |
| T | 6706 hCoV-19/Australia/VIC17043/2020\|EPI_ISL_663700\|202 | ···················· | ···················· | ···················· | ···················· | ···················· | ···················· |
| T | 6707 hCoV-19/Australia/VIC17044/2020\|EPI_ISL_663701\|202 | ···················· | ···················· | ···················· | ···················· | ···················· | ···················· |
| T | 6708 hCoV-19/Australia/VIC17045/2020\|EPI_ISL_663702\|202 | ···················· | ···················· | ···················· | ···················· | ···················· | ···················· |
| T | 6709 hCoV-19/Australia/VIC17047/2020\|EPI_ISL_663703\|202 | ···················· | ···················· | ···················· | ···················· | ···················· | ···················· |
| T | 6710 hCoV-19/Australia/VIC17048/2020\|EPI_ISL_663704\|202 | ···················· | ···················· | ···················· | ···················· | ···················· | ···················· |
| T | 6711 hCoV-19/Australia/VIC17049/2020\|EPI_ISL_663705\|202 | ···················· | ···················· | ···················· | ···················· | ···················· | ···················· |
| T | 6712 hCoV-19/Australia/VIC17050/2020\|EPI_ISL_663706\|202 | ···················· | ···················· | ···················· | ···················· | ···················· | ···················· |
| T | 6713 hCoV-19/Australia/VIC17051/2020\|EPI_ISL_663707\|202 | ···················· | ···················· | ···················· | ···················· | ···················· | ···················· |
| T | 6714 hCoV-19/Australia/VIC17052/2020\|EPI_ISL_663708\|202 | ···················· | ···················· | ···················· | ···················· | ···················· | ···················· |
| T | 6715 hCoV-19/Australia/VIC17053/2020\|EPI_ISL_663709\|202 | ···················· | ···················· | ···················· | ···················· | ···················· | ···················· |
| T | 6716 hCoV-19/Australia/VIC17056/2020\|EPI_ISL_663710\|202 | ···················· | ···················· | ···················· | ···················· | ···················· | ···················· |
| T | 6717 hCoV-19/Australia/VIC17057/2020\|EPI_ISL_663711\|202 | ···················· | ···················· | ···················· | ···················· | ···················· | ···················· |
| T | 6718 hCoV-19/Australia/VIC17060/2020\|EPI_ISL_663712\|202 | ···················· | ···················· | ···················· | ···················· | ···················· | ···················· |
| T | 6719 hCoV-19/Australia/VIC17066/2020\|EPI_ISL_663713\|202 | ···················· | ···················· | ···················· | ···················· | ···················· | ···················· |
| T | 6720 hCoV-19/Australia/VIC17067/2020\|EPI_ISL_663714\|202 | ···················· | ···················· | ···················· | ···················· | ···················· | ···················· |
| T | 6721 hCoV-19/Australia/VIC17068/2020\|EPI_ISL_663715\|202 | ···················· | ···················· | ···················· | ···················· | ···················· | ···················· |
| T | 6722 hCoV-19/Australia/VIC17070/2020\|EPI_ISL_663716\|202 | ···················· | ···················· | ···················· | ···················· | ···················· | ···················· |
| T | 6723 hCoV-19/Australia/VIC17071/2020\|EPI_ISL_663717\|202 | ···················· | ···················· | ···················· | ···················· | ···················· | ···················· |

```
T    6724 hCoV-19/Australia/VIC17075/2020|EPI_ISL_663718|202   ····················  ····················  ····················  ····················  ····················  ····················
T    6725 hCoV-19/Australia/VIC17078/2020|EPI_ISL_663719|202   ····················  ····················  ····················  ····················  ····················  ····················
T    6726 hCoV-19/Australia/VIC17080/2020|EPI_ISL_663720|202   ····················  ····················  ····················  ····················  ····················  ····················
T    6727 hCoV-19/Australia/VIC17081/2020|EPI_ISL_663721|202   ····················  ····················  ····················  ····················  ····················  ····················
T    6728 hCoV-19/Australia/VIC17082/2020|EPI_ISL_663722|202   ····················  ····················  ····················  ····················  ····················  ····················
T    6729 hCoV-19/Australia/VIC17085/2020|EPI_ISL_663723|202   ····················  ····················  ····················  ····················  ····················  ····················
T    6730 hCoV-19/Australia/VIC17086/2020|EPI_ISL_663724|202   ····················  ····················  ····················  ····················  ····················  ····················
T    6731 hCoV-19/Australia/VIC17088/2020|EPI_ISL_663725|202   ····················  ····················  ····················  ····················  ····················  ····················
T    6732 hCoV-19/Australia/VIC17091/2020|EPI_ISL_663726|202   ····················  ····················  ····················  ····················  ····················  ····················
T    6733 hCoV-19/Australia/VIC17092/2020|EPI_ISL_663727|202   ····················  ····················  ····················  ····················  ····················  ····················
T    6734 hCoV-19/Australia/VIC17094/2020|EPI_ISL_663728|202   ····················  ····················  ····················  ····················  ····················  ····················
T    6735 hCoV-19/Australia/VIC17097/2020|EPI_ISL_663729|202   ····················  ····················  ····················  ····················  ····················  ····················
T    6736 hCoV-19/Australia/VIC17098/2020|EPI_ISL_663730|202   ····················  ····················  ····················  ····················  ····················  ····················
T    6737 hCoV-19/Australia/VIC17099/2020|EPI_ISL_663731|202   ····················  ····················  ····················  ····················  ····················  ····················
T    6738 hCoV-19/Australia/VIC17101/2020|EPI_ISL_663732|202   ····················  ····················  ····················  ····················  ····················  ····················
T    6739 hCoV-19/Australia/VIC17103/2020|EPI_ISL_663733|202   ····················  ····················  ····················  ····················  ····················  ····················
T    6740 hCoV-19/Australia/VIC17105/2020|EPI_ISL_663734|202   ····················  ····················  ····················  ····················  ····················  ····················
T    6741 hCoV-19/Australia/VIC17106/2020|EPI_ISL_663735|202   ····················  ····················  ····················  ····················  ····················  ····················
T    6742 hCoV-19/Australia/VIC17107/2020|EPI_ISL_663736|202   ····················  ····················  ····················  ····················  ····················  ····················
T    6743 hCoV-19/Australia/VIC17109/2020|EPI_ISL_663737|202   ····················  ····················  ····················  ····················  ····················  ····················
T    6744 hCoV-19/Australia/VIC17113/2020|EPI_ISL_663738|202   ····················  ····················  ····················  ····················  ····················  ····················
T    6745 hCoV-19/Australia/VIC17114/2020|EPI_ISL_663739|202   ····················  ····················  ····················  ····················  ····················  ····················
T    6746 hCoV-19/Australia/VIC17116/2020|EPI_ISL_663740|202   ····················  ····················  ····················  ····················  ····················  ····················
T    6747 hCoV-19/Australia/VIC17117/2020|EPI_ISL_663741|202   ····················  ····················  ····················  ····················  ····················  ····················
T    6748 hCoV-19/Australia/VIC17149/2020|EPI_ISL_663765|202   ····················  ····················  ····················  ····················  ····················  ····················
T    6749 hCoV-19/Australia/VIC17164/2020|EPI_ISL_663775|202   ····················  ····················  ····················  ····················  ····················  ····················
T    6750 hCoV-19/Australia/VIC17165/2020|EPI_ISL_663776|202   ····················  ····················  ····················  ····················  ····················  ····················
T    6751 hCoV-19/Australia/VIC17167/2020|EPI_ISL_663777|202   ····················  ····················  ····················  ····················  ····················  ····················
T    6752 hCoV-19/Australia/VIC17171/2020|EPI_ISL_663778|202   ····················  ····················  ····················  ····················  ····················  ····················
T    6756 hCoV-19/Australia/VIC17184/2020|EPI_ISL_663782|202   ····················  ····················  ····················  ····················  ····················  ····················
T    6758 hCoV-19/Australia/VIC17190/2020|EPI_ISL_663784|202   ····················  ····················  ····················  ····················  ····················  ····················
T    6759 hCoV-19/Australia/VIC17192/2020|EPI_ISL_663785|202   ····················  ····················  ····················  ····················  ····················  ····················
T    6760 hCoV-19/Australia/VIC17193/2020|EPI_ISL_663786|202   ····················  ····················  ····················  ····················  ····················  ····················
T    6761 hCoV-19/Australia/VIC17194/2020|EPI_ISL_663787|202   ····················  ····················  ····················  ····················  ····················  ····················
T    6762 hCoV-19/Australia/VIC17195/2020|EPI_ISL_663788|202   ····················  ····················  ····················  ····················  ····················  ····················
T    6763 hCoV-19/Australia/VIC17198/2020|EPI_ISL_663789|202   ····················  ····················  ····················  ····················  ····················  ····················
T    6764 hCoV-19/Australia/VIC17201/2020|EPI_ISL_663790|202   ····················  ····················  ····················  ····················  ····················  ····················
T    6765 hCoV-19/Australia/VIC17202/2020|EPI_ISL_663791|202   ····················  ····················  ····················  ····················  ····················  ····················
T    6766 hCoV-19/Australia/VIC17203/2020|EPI_ISL_663792|202   ····················  ····················  ····················  ····················  ····················  ····················
T    6767 hCoV-19/Australia/VIC17204/2020|EPI_ISL_663793|202   ····················  ····················  ····················  ····················  ····················  ····················
T    6768 hCoV-19/Australia/VIC17206/2020|EPI_ISL_663794|202   ····················  ····················  ····················  ····················  ····················  ····················
T    6769 hCoV-19/Australia/VIC17208/2020|EPI_ISL_663795|202   ····················  ····················  ····················  ····················  ····················  ····················
T    6770 hCoV-19/Australia/VIC17210/2020|EPI_ISL_663796|202   ····················  ····················  ····················  ····················  ····················  ····················
T    6771 hCoV-19/Australia/VIC17215/2020|EPI_ISL_663797|202   ····················  ····················  ····················  ····················  ····················  ····················
T    6772 hCoV-19/Australia/VIC17216/2020|EPI_ISL_663798|202   ····················  ····················  ····················  ····················  ····················  ····················
T    6773 hCoV-19/Australia/VIC17218/2020|EPI_ISL_663799|202   ····················  ····················  ····················  ····················  ····················  ····················
T    6774 hCoV-19/England/QEUH-AA0493/2020|EPI_ISL_666000|20   ····················  ····················  ····················  ····················  ····················  ····················
T    6775 hCoV-19/England/QEUH-AA04A2/2020|EPI_ISL_666001|20   ····················  ····················  ····················  ····················  ····················  ····················
T    6776 hCoV-19/England/QEUH-AA06BB/2020|EPI_ISL_666002|20   ····················  ····················  ····················  ····················  ····················  ····················
T    6777 hCoV-19/England/QEUH-AF0D4E/2020|EPI_ISL_666003|20   ····················  ····················  ····················  ····················  ····················  ····················
T    6778 hCoV-19/England/QEUH-AF0D99/2020|EPI_ISL_666004|20   ····················  ·A··················  ····················  ····················  ····················  ··A·················
T    6779 hCoV-19/England/QEUH-AF0DA8/2020|EPI_ISL_666005|20   ····················  ····················  ····················  ····················  ····················  ····················
T    6780 hCoV-19/England/QEUH-AF0DD5/2020|EPI_ISL_666006|20   ····················  ····················  ····················  ····················  ····················  ····················
T    6781 hCoV-19/England/QEUH-AF119C/2020|EPI_ISL_666007|20   ····················  ····················  ····················  ····················  ····················  ····················
T    6782 hCoV-19/England/QEUH-AF14B1/2020|EPI_ISL_666012|20   ····················  ····················  ····················  ····················  ····················  ····················
T    6783 hCoV-19/England/QEUH-AF15DC/2020|EPI_ISL_666015|20   ····················  ····················  ····················  ····················  ····················  ····················
T    6784 hCoV-19/England/QEUH-AF15EB/2020|EPI_ISL_666016|20   ····················  ····················  ····················  ····················  ····················  ····················
T    6785 hCoV-19/England/QEUH-AF1615/2020|EPI_ISL_666017|20   ····················  ····················  ····················  ····················  ····················  ····················
T    6786 hCoV-19/England/QEUH-AF18C4/2020|EPI_ISL_666018|20   ····················  ····················  ····················  ····················  ····················  ····················
T    6787 hCoV-19/USA/OR-OHSU-2639/2020|EPI_ISL_667270|2020-   ····················  ····················  ····················  ····················  ····················  ····················
T    6788 hCoV-19/USA/OR-OHSU-2641/2020|EPI_ISL_667271|2020-   ····················  ·A··················  ····················  ····················  ····················  ····················
T    6789 hCoV-19/USA/OR-OHSU-2649/2020|EPI_ISL_667273|2020-   ····················  ·A··················  ····················  ····················  ····················  ····················
T    6790 hCoV-19/USA/OR-OHSU-2655/2020|EPI_ISL_667274|2020-   ····················  ····················  ····················  ····················  ····················  ····················
T    6791 hCoV-19/USA/NJ-OHSU-2657/2020|EPI_ISL_667275|2020-   ····················  ·A··················  ····················  ····················  ····················  ····················
T    6792 hCoV-19/USA/OR-OHSU-2661/2020|EPI_ISL_667276|2020-   ····················  ····················  ····················  ····················  ····················  ····················
T    6793 hCoV-19/USA/OR-OHSU-2667/2020|EPI_ISL_667277|2020-   ····················  ·A··················  ····················  ····················  ····················  ····················
T    6794 hCoV-19/USA/OR-OHSU-2675/2020|EPI_ISL_667278|2020-   ····················  ····················  ····················  ····················  ····················  ····················
T    6795 hCoV-19/USA/OR-OHSU-2681/2020|EPI_ISL_667280|2020-   ····················  ·A··················  ····················  ····················  ····················  ····················
T    6796 hCoV-19/USA/OR-OHSU-2687/2020|EPI_ISL_667282|2020-   ····················  ····················  ····················  ····················  ····················  ····················
T    6797 hCoV-19/USA/OR-OHSU-2689/2020|EPI_ISL_667283|2020-   ····················  ····················  ····················  ····················  ····················  ····················
T    6798 hCoV-19/USA/OR-OHSU-2695/2020|EPI_ISL_667285|2020-   ····················  ····················  ····················  ····················  ····················  ····················
T    6799 hCoV-19/USA/OR-OHSU-2697/2020|EPI_ISL_667286|2020-   ····················  ····················  ····················  ····················  ····················  ····················
T    6800 hCoV-19/USA/OR-OHSU-2707/2020|EPI_ISL_667289|2020-   ····················  ····················  ····················  ····················  ····················  ····················
T    6801 hCoV-19/USA/OR-OHSU-2711/2020|EPI_ISL_667291|2020-   ····················  ····················  ····················  ····················  ····················  ····················
T    6802 hCoV-19/USA/OR-OHSU-2715/2020|EPI_ISL_667292|2020-   ····················  ····················  ····················  ····················  ····················  ····················
T    6803 hCoV-19/USA/WA-OHSU-2719/2020|EPI_ISL_667293|2020-   ····················  ····················  ····················  ····················  ····················  ····················
T    6804 hCoV-19/USA/OR-OHSU-2723/2020|EPI_ISL_667294|2020-   ····················  ····················  ····················  ····················  ····················  ····················
T    6805 hCoV-19/USA/OR-OHSU-2735/2020|EPI_ISL_667296|2020-   ····················  ·A··················  ····················  ····················  ····················  ····················
T    6806 hCoV-19/USA/OR-OHSU-2737/2020|EPI_ISL_667297|2020-   ····················  ····················  ····················  ····················  ····················  ····················
T    6807 hCoV-19/USA/OR-OHSU-2741/2020|EPI_ISL_667298|2020-   ····················  ····················  ····················  ····················  ····················  ····················
```

| | | | | | | | | |
|---|---|---|---|---|---|---|---|---|
| T | 6808 | hCoV-19/USA/OR-OHSU-2743/2020\|EPI_ISL_667299\|2020- | ···················· | ···················· | ····················· | ···················· | ···················· | ···················· |
| T | 6809 | hCoV-19/USA/OR-OHSU-2771/2020\|EPI_ISL_667307\|2020- | ···················· | ···················· | ····················· | ···················· | ···················· | ···················· |
| T | 6810 | hCoV-19/USA/OR-OHSU-2773/2020\|EPI_ISL_667308\|2020- | ···················· | ···················· | ····················· | ···················· | ···················· | ···················· |
| T | 6811 | hCoV-19/USA/OR-OHSU-2775/2020\|EPI_ISL_667309\|2020- | ···················· | ···················· | ····················· | ···················· | ···················· | ···················· |
| T | 6812 | hCoV-19/USA/OR-OHSU-2777/2020\|EPI_ISL_667310\|2020- | ···················· | ········A··········· | ····················· | ···················· | ···················· | ···················· |
| T | 6813 | hCoV-19/USA/OR-OHSU-2783/2020\|EPI_ISL_667311\|2020- | ···················· | ·A·················· | ····················· | ···················· | ···················· | ···················· |
| T | 6814 | hCoV-19/USA/OR-OHSU-2791/2020\|EPI_ISL_667313\|2020- | ···················· | ···················· | ····················· | ···················· | ···················· | ···················· |
| T | 6815 | hCoV-19/USA/OR-OHSU-2797/2020\|EPI_ISL_667315\|2020- | ···················· | ···················· | ····················· | ···················· | ···················· | ···················· |
| T | 6816 | hCoV-19/USA/OR-OHSU-2799/2020\|EPI_ISL_667316\|2020- | ···················· | ·A·················· | ····················· | ···················· | ···················· | ···················· |
| T | 6817 | hCoV-19/USA/OR-OHSU-2803/2020\|EPI_ISL_667317\|2020- | ···················· | ···················· | ····················· | ···················· | ···················· | ···················· |
| T | 6818 | hCoV-19/USA/OR-OHSU-2810/2020\|EPI_ISL_667319\|2020- | ···················· | ·A·················· | ····················· | ···················· | ···················· | ···················· |
| T | 6819 | hCoV-19/USA/OR-OHSU-2811/2020\|EPI_ISL_667320\|2020- | ···················· | ·A·················· | ····················· | ···················· | ···················· | ···················· |
| T | 6820 | hCoV-19/USA/OR-OHSU-2812/2020\|EPI_ISL_667321\|2020- | ···················· | ···················· | ····················· | ···················· | ···················· | ···················· |
| T | 6821 | hCoV-19/USA/OR-OHSU-2813/2020\|EPI_ISL_667322\|2020- | ···················· | ·A·················· | ····················· | ···················· | ···················· | ···················· |
| T | 6822 | hCoV-19/USA/OR-OHSU-2814/2020\|EPI_ISL_667323\|2020- | ···················· | ···················· | ····················· | ···················· | ···················· | ···················· |
| T | 6823 | hCoV-19/USA/OR-OHSU-2815/2020\|EPI_ISL_667324\|2020- | ···················· | ···················· | ····················· | ···················· | ···················· | ···················· |
| T | 6824 | hCoV-19/USA/OR-OHSU-2817/2020\|EPI_ISL_667325\|2020- | ···················· | ···················· | ····················· | ···················· | ···················· | ···················· |
| T | 6825 | hCoV-19/USA/OR-OHSU-2819/2020\|EPI_ISL_667327\|2020- | ···················· | ···················· | ····················· | ···················· | ···················· | ···················· |
| T | 6826 | hCoV-19/USA/OR-OHSU-2820/2020\|EPI_ISL_667328\|2020- | ···················· | ·A·················· | ····················· | ···················· | ···················· | ···················· |
| T | 6827 | hCoV-19/USA/OR-OHSU-2821/2020\|EPI_ISL_667329\|2020- | ···················· | ·A·················· | ····················· | ···················· | ···················· | ···················· |
| T | 6828 | hCoV-19/USA/OR-OHSU-2822/2020\|EPI_ISL_667330\|2020- | ········T··········· | ···················· | ····················· | ···················· | ···················· | ···················· |
| T | 6829 | hCoV-19/USA/OR-OHSU-2823/2020\|EPI_ISL_667331\|2020- | ···················· | ·A·················· | ····················· | ···················· | ···················· | ···················· |
| T | 6830 | hCoV-19/USA/OR-OHSU-2824/2020\|EPI_ISL_667332\|2020- | ···················· | ···················· | ····················· | ···················· | ···················· | ···················· |
| T | 6831 | hCoV-19/USA/OR-OHSU-2826/2020\|EPI_ISL_667334\|2020- | ···················· | ···················· | ····················· | ···················· | ···················· | ···················· |
| T | 6832 | hCoV-19/USA/OR-OHSU-2827/2020\|EPI_ISL_667335\|2020- | ···················· | ·A·················· | ····················· | ···················· | ···················· | ···················· |
| T | 6833 | hCoV-19/USA/OR-OHSU-2831/2020\|EPI_ISL_667338\|2020- | ···················· | ·A·················· | ····················· | ···················· | ···················· | ···················· |
| T | 6834 | hCoV-19/USA/OR-OHSU-2842/2020\|EPI_ISL_667347\|2020- | ···················· | ···················· | ····················· | ···················· | ···················· | ···················· |
| T | 6835 | hCoV-19/USA/OR-OHSU-2843/2020\|EPI_ISL_667348\|2020- | ···················· | ···················· | ····················· | ···················· | ···················· | ···················· |
| T | 6836 | hCoV-19/Denmark/DCGC-9516/2020\|EPI_ISL_668637\|2020 | ···················· | ···················· | ····················· | ···················· | ···················· | ···················· |
| T | 6837 | hCoV-19/Denmark/DCGC-9624/2020\|EPI_ISL_668638\|2020 | ···················· | ···················· | ····················· | ···················· | ···················· | ···················· |
| T | 6838 | hCoV-19/Denmark/DCGC-10091/2020\|EPI_ISL_668639\|202 | ···················· | ···················· | ····················· | ···················· | ···················· | ···················· |
| T | 6839 | hCoV-19/Denmark/DCGC-10185/2020\|EPI_ISL_668640\|202 | ···················· | ···················· | ····················· | ···················· | ···················· | ···················· |
| T | 6840 | hCoV-19/Denmark/DCGC-10200/2020\|EPI_ISL_668641\|202 | ···················· | ···················· | ····················· | ···················· | ···················· | ···················· |
| T | 6841 | hCoV-19/Denmark/DCGC-9250/2020\|EPI_ISL_668663\|2020 | ···················· | ···················· | ····················· | ···················· | ···················· | ···················· |
| T | 6842 | hCoV-19/Denmark/DCGC-9256/2020\|EPI_ISL_668664\|2020 | ···················· | ···················· | ····················· | ···················· | ···················· | ···················· |
| T | 6843 | hCoV-19/Denmark/DCGC-9263/2020\|EPI_ISL_668665\|2020 | ···················· | ···················· | ····················· | ···················· | ···················· | ···················· |
| T | 6844 | hCoV-19/Denmark/DCGC-9265/2020\|EPI_ISL_668666\|2020 | ···················· | ···················· | ····················· | ···················· | ···················· | ···················· |
| T | 6845 | hCoV-19/Denmark/DCGC-9267/2020\|EPI_ISL_668667\|2020 | ···················· | ···················· | ····················· | ···················· | ···················· | ···················· |
| T | 6846 | hCoV-19/Denmark/DCGC-9274/2020\|EPI_ISL_668668\|2020 | ···················· | ···················· | ····················· | ···················· | ···················· | ···················· |
| T | 6847 | hCoV-19/Denmark/DCGC-9285/2020\|EPI_ISL_668669\|2020 | ···················· | ···················· | ····················· | ···················· | ···················· | ···················· |
| T | 6848 | hCoV-19/Denmark/DCGC-9463/2020\|EPI_ISL_668677\|2020 | ···················· | ···················· | ····················· | ···················· | ···················· | ···················· |
| T | 6849 | hCoV-19/Denmark/DCGC-9465/2020\|EPI_ISL_668678\|2020 | ···················· | ···················· | ····················· | ···················· | ···················· | ···················· |
| T | 6850 | hCoV-19/Denmark/DCGC-9509/2020\|EPI_ISL_668679\|2020 | ···················· | ···················· | ····················· | ···················· | ···················· | ···················· |
| T | 6851 | hCoV-19/Denmark/DCGC-10435/2020\|EPI_ISL_669968\|202 | ···················· | ···················· | ····················· | ···················· | ···················· | ···················· |
| T | 6852 | hCoV-19/Denmark/DCGC-10437/2020\|EPI_ISL_669969\|202 | ···················· | ···················· | ····················· | ···················· | ···················· | ···················· |
| T | 6853 | hCoV-19/Denmark/DCGC-10438/2020\|EPI_ISL_669970\|202 | ···················· | ···················· | ····················· | ···················· | ···················· | ···················· |
| T | 6854 | hCoV-19/Denmark/DCGC-10440/2020\|EPI_ISL_669971\|202 | ···················· | ···················· | ····················· | ···················· | ···················· | ···················· |
| T | 6855 | hCoV-19/Denmark/DCGC-10441/2020\|EPI_ISL_669972\|202 | ···················· | ···················· | ····················· | ···················· | ···················· | ···················· |
| T | 6856 | hCoV-19/Denmark/DCGC-10442/2020\|EPI_ISL_669973\|202 | ···················· | ···················· | ····················· | ···················· | ···················· | ···················· |
| T | 6857 | hCoV-19/Denmark/DCGC-10443/2020\|EPI_ISL_669974\|202 | ···················· | ···················· | ····················· | ···················· | ···················· | ···················· |
| T | 6858 | hCoV-19/Denmark/DCGC-10444/2020\|EPI_ISL_669975\|202 | ···················· | ···················· | ····················· | ···················· | ···················· | ···················· |
| T | 6859 | hCoV-19/Denmark/DCGC-10445/2020\|EPI_ISL_669976\|202 | ···················· | ···················· | ····················· | ···················· | ···················· | ···················· |
| T | 6860 | hCoV-19/Denmark/DCGC-10446/2020\|EPI_ISL_669977\|202 | ···················· | ···················· | ····················· | ···················· | ···················· | ···················· |
| T | 6861 | hCoV-19/Denmark/DCGC-10447/2020\|EPI_ISL_669978\|202 | ···················· | ·A·················· | ····················· | ···················· | ···················· | ···················· |
| T | 6862 | hCoV-19/Denmark/DCGC-10448/2020\|EPI_ISL_669979\|202 | ···················· | ···················· | ····················· | ···················· | ···················· | ···················· |
| T | 6863 | hCoV-19/Denmark/DCGC-10449/2020\|EPI_ISL_669980\|202 | ···················· | ···················· | ····················· | ···················· | ···················· | ···················· |
| T | 6864 | hCoV-19/Denmark/DCGC-10451/2020\|EPI_ISL_669981\|202 | ···················· | ···················· | ····················· | ···················· | ···················· | ···················· |
| T | 6865 | hCoV-19/Denmark/DCGC-10452/2020\|EPI_ISL_669982\|202 | ···················· | ···················· | ····················· | ···················· | ···················· | ···················· |
| T | 6866 | hCoV-19/Denmark/DCGC-10453/2020\|EPI_ISL_669983\|202 | ···················· | ···················· | ····················· | ···················· | ···················· | ···················· |
| T | 6867 | hCoV-19/Denmark/DCGC-10454/2020\|EPI_ISL_669984\|202 | ···················· | ···················· | ····················· | ···················· | ···················· | ···················· |
| T | 6868 | hCoV-19/Denmark/DCGC-10455/2020\|EPI_ISL_669985\|202 | ···················· | ···················· | ····················· | ···················· | ···················· | ···················· |
| T | 6869 | hCoV-19/Denmark/DCGC-10456/2020\|EPI_ISL_669986\|202 | ···················· | ···················· | ····················· | ···················· | ···················· | ···················· |
| T | 6870 | hCoV-19/Denmark/DCGC-10458/2020\|EPI_ISL_669987\|202 | ···················· | ···················· | ····················· | ···················· | ···················· | ···················· |
| T | 6871 | hCoV-19/Denmark/DCGC-10459/2020\|EPI_ISL_669988\|202 | ···················· | ···················· | ····················· | ···················· | ···················· | ···················· |
| T | 6872 | hCoV-19/Denmark/DCGC-10461/2020\|EPI_ISL_669989\|202 | ···················· | ···················· | ····················· | ···················· | ···················· | ···················· |
| T | 6873 | hCoV-19/Denmark/DCGC-10462/2020\|EPI_ISL_669990\|202 | ···················· | ···················· | ····················· | ···················· | ···················· | ···················· |
| T | 6874 | hCoV-19/Denmark/DCGC-10463/2020\|EPI_ISL_669991\|202 | ···················· | ···················· | ····················· | ···················· | ···················· | ···················· |
| T | 6875 | hCoV-19/Denmark/DCGC-10465/2020\|EPI_ISL_669992\|202 | ···················· | ···················· | ····················· | ···················· | ···················· | ···················· |
| T | 6876 | hCoV-19/Denmark/DCGC-10466/2020\|EPI_ISL_669993\|202 | ···················· | ···················· | ····················· | ···················· | ···················· | ···················· |
| T | 6877 | hCoV-19/Denmark/DCGC-10467/2020\|EPI_ISL_669994\|202 | ···················· | ···················· | ····················· | ···················· | ···················· | ···················· |
| T | 6878 | hCoV-19/Denmark/DCGC-10468/2020\|EPI_ISL_669995\|202 | ···················· | ···················· | ····················· | ···················· | ···················· | ···················· |
| T | 6879 | hCoV-19/Denmark/DCGC-10471/2020\|EPI_ISL_669996\|202 | ···················· | ···················· | ····················· | ···················· | ···················· | ···················· |
| T | 6880 | hCoV-19/Denmark/DCGC-10473/2020\|EPI_ISL_669997\|202 | ···················· | ···················· | ····················· | ···················· | ···················· | ···················· |
| T | 6881 | hCoV-19/Denmark/DCGC-10475/2020\|EPI_ISL_669998\|202 | ···················· | ···················· | ····················· | ···················· | ···················· | ···················· |
| T | 6882 | hCoV-19/Denmark/DCGC-10476/2020\|EPI_ISL_669999\|202 | ···················· | ···················· | ····················· | ···················· | ···················· | ···················· |
| T | 6883 | hCoV-19/Denmark/DCGC-11486/2020\|EPI_ISL_670950\|202 | ···················· | ·A·················· | ····················· | ···················· | ···················· | ···A················ |
| T | 6884 | hCoV-19/Denmark/DCGC-11487/2020\|EPI_ISL_670951\|202 | ···················· | ···················· | ····················· | ···················· | ···················· | ···················· |
| T | 6885 | hCoV-19/Denmark/DCGC-11488/2020\|EPI_ISL_670952\|202 | ···················· | ·A·················· | ····················· | ···················· | ···················· | ···A················ |
| T | 6886 | hCoV-19/Denmark/DCGC-11489/2020\|EPI_ISL_670953\|202 | ···················· | ···················· | ····················· | ···················· | ···················· | ···················· |
| T | 6887 | hCoV-19/Denmark/DCGC-11491/2020\|EPI_ISL_670954\|202 | ···················· | ···················· | ····················· | ···················· | ···················· | ···················· |

T 6888 hCoV-19/Denmark/DCGC-11492/2020|EPI_ISL_670955|202 ···················· ···················· ····················· ···················· ···················· ····················
T 6889 hCoV-19/Denmark/DCGC-11493/2020|EPI_ISL_670956|202 ···················· ···················· ····················· ···················· ···················· ····················
T 6890 hCoV-19/Denmark/DCGC-11495/2020|EPI_ISL_670957|202 ···················· ···················· ····················· ···················· ···················· ····················
T 6891 hCoV-19/Denmark/DCGC-11496/2020|EPI_ISL_670958|202 ···················· ···················· ····················· ···················· ···················· ····················
T 6892 hCoV-19/Denmark/DCGC-11497/2020|EPI_ISL_670959|202 ···················· ···················· ····················· ···················· ···················· ····················
T 6893 hCoV-19/Denmark/DCGC-11500/2020|EPI_ISL_670960|202 ···················· ···················· ····················· ···················· ···················· ····················
T 6894 hCoV-19/Denmark/DCGC-11501/2020|EPI_ISL_670961|202 ···················· ···················· ····················· ···················· ···················· ····················
T 6895 hCoV-19/Denmark/DCGC-11504/2020|EPI_ISL_670962|202 ···················· ···················· ····················· ···················· ···················· ····················
T 6896 hCoV-19/Denmark/DCGC-11506/2020|EPI_ISL_670963|202 ···················· ···················· ····················· ···················· ···················· ····················
T 6897 hCoV-19/Denmark/DCGC-11508/2020|EPI_ISL_670964|202 ···················· ·A·················· ····················· ···················· ···················· ····················
T 6898 hCoV-19/Denmark/DCGC-11509/2020|EPI_ISL_670965|202 ···················· ···················· ····················· ···················· ···················· ····················
T 6899 hCoV-19/Denmark/DCGC-11510/2020|EPI_ISL_670966|202 ···················· ···················· ····················· ···················· ···················· ····················
T 6900 hCoV-19/Denmark/DCGC-11512/2020|EPI_ISL_670967|202 ···················· ·A···A·············· ····················· ···················· ···················· ····················
T 6901 hCoV-19/Denmark/DCGC-11513/2020|EPI_ISL_670968|202 ···················· ·A·················· ····················· ···················· ···················· ····················
T 6902 hCoV-19/Denmark/DCGC-11514/2020|EPI_ISL_670969|202 ···················· ···················· ····················· ···················· ···················· ····················
T 6903 hCoV-19/Denmark/DCGC-11515/2020|EPI_ISL_670970|202 ···················· ···················· ····················· ···················· ···················· ····················
T 6904 hCoV-19/Denmark/DCGC-11516/2020|EPI_ISL_670971|202 ···················· ···················· ····················· ···················· ···················· ····················
T 6905 hCoV-19/Denmark/DCGC-11518/2020|EPI_ISL_670972|202 ···················· ···················· ····················· ···················· ···················· ····················
T 6906 hCoV-19/Denmark/DCGC-11519/2020|EPI_ISL_670973|202 ···················· ···················· ····················· ···················· ···················· ····················
T 6907 hCoV-19/Denmark/DCGC-11523/2020|EPI_ISL_670974|202 ···················· ···················· ····················· ···················· ···················· ····················
T 6908 hCoV-19/Denmark/DCGC-11524/2020|EPI_ISL_670975|202 ···················· ···················· ····················· ···················· ···················· ····················
T 6909 hCoV-19/Denmark/DCGC-11528/2020|EPI_ISL_670976|202 ···················· ···················· ····················· ···················· ···················· ····················
T 6910 hCoV-19/Denmark/DCGC-11530/2020|EPI_ISL_670977|202 ···················· ·A·················· ····················· ···················· ···················· ····················
T 6911 hCoV-19/Denmark/DCGC-11532/2020|EPI_ISL_670978|202 ···················· ···················· ····················· ···················· ···················· ····················
T 6912 hCoV-19/Denmark/DCGC-11533/2020|EPI_ISL_670979|202 ···················· ···················· ····················· ···················· ···················· ····················
T 6913 hCoV-19/England/QEUH-B6B150/2020|EPI_ISL_662378|20 ···················· ···················· ····················· ···················· ···················· ····················
T 6914 hCoV-19/England/QEUH-B6A79B/2020|EPI_ISL_662379|20 ···················· ···················· ····················· ···················· ···················· ····················
T 6915 hCoV-19/England/QEUH-B6A3C4/2020|EPI_ISL_662380|20 ···················· ···················· ····················· ···················· ···················· ····················
T 6916 hCoV-19/England/ALDP-B73692/2020|EPI_ISL_662381|20 ···················· ···················· ····················· ···················· ···················· ··A·················
T 6917 hCoV-19/England/ALDP-B7346B/2020|EPI_ISL_662382|20 ···················· ···················· ····················· ···················· ···················· ····················
T 6918 hCoV-19/England/QEUH-B6B105/2020|EPI_ISL_662383|20 ···················· ···················· ····················· ···················· ···················· ····················
T 6919 hCoV-19/England/QEUH-B6A142/2020|EPI_ISL_662384|20 ···················· ···················· ····················· ···················· ···················· ····················
T 6920 hCoV-19/England/ALDP-B73726/2020|EPI_ISL_662385|20 ···················· ···················· ····················· ···················· ···················· ····················
T 6921 hCoV-19/England/QEUH-B6A986/2020|EPI_ISL_662386|20 ···················· ···················· ····················· ···················· ···················· ····················
T 6922 hCoV-19/England/QEUH-B6A379/2020|EPI_ISL_662387|20 ···················· ···················· ····················· ···················· ···················· ····················
T 6923 hCoV-19/England/QEUH-B6A124/2020|EPI_ISL_662388|20 ···················· ···················· ····················· ···················· ···················· ····················
T 6924 hCoV-19/England/ALDP-B736DE/2020|EPI_ISL_662389|20 ···················· ···················· ····················· ···················· ···················· ····················
T 6925 hCoV-19/England/ALDP-B73498/2020|EPI_ISL_662390|20 ···················· ···················· ····················· ···················· ···················· ····················
T 6926 hCoV-19/England/ALDP-B735B3/2020|EPI_ISL_662391|20 ···················· ···················· ····················· ···················· ···················· ····················
T 6927 hCoV-19/England/QEUH-B66AF0/2020|EPI_ISL_662392|20 ···················· ···················· ····················· ···················· ···················· ····················
T 6928 hCoV-19/England/ALDP-B73717/2020|EPI_ISL_662393|20 ···················· ···················· ····················· ···················· ···················· ····················
T 6929 hCoV-19/England/QEUH-B6AE3C/2020|EPI_ISL_662395|20 ···················· ···················· ····················· ···················· ···················· ····················
T 6930 hCoV-19/England/QEUH-B672D9/2020|EPI_ISL_662396|20 ···················· ···················· ····················· ···················· ···················· ··A·················
T 6931 hCoV-19/England/QEUH-B66B1B/2020|EPI_ISL_662399|20 ···················· ···················· ····················· ···················· ···················· ····················
T 6932 hCoV-19/England/ALDP-B733B9/2020|EPI_ISL_662411|20 ···················· ···················· ····················· ···················· ···················· ····················
T 6933 hCoV-19/England/QEUH-B6A1BB/2020|EPI_ISL_662414|20 ···················· ···················· ····················· ···················· ···················· ····················
T 6934 hCoV-19/England/ALDP-B7361A/2020|EPI_ISL_662415|20 ···················· ·A·················· ····················· ···················· ···················· ····················
T 6935 hCoV-19/England/QEUH-B6A41C/2020|EPI_ISL_662416|20 ···················· ···················· ····················· ···················· ···················· ····················
T 6936 hCoV-19/England/QEUH-B66410/2020|EPI_ISL_662417|20 ···················· ···················· ····················· ···················· ···················· ····················
T 6937 hCoV-19/England/ALDP-B73638/2020|EPI_ISL_662419|20 ···················· ···················· ····················· ···················· ···················· ····················
T 6938 hCoV-19/England/QEUH-B66B66/2020|EPI_ISL_662420|20 ···················· ···················· ····················· ···················· ···················· ····················
T 6939 hCoV-19/England/QEUH-B6686F/2020|EPI_ISL_662421|20 ···················· ···················· ····················· ···················· ···················· ····················
T 6940 hCoV-19/England/QEUH-B6A19D/2020|EPI_ISL_662422|20 ···················· ···················· ····················· ···················· ···················· ····················
T 6941 hCoV-19/England/QEUH-B66BEE/2020|EPI_ISL_662423|20 ···················· ···················· ····················· ···················· ···················· ····················
T 6942 hCoV-19/England/QEUH-B674F1/2020|EPI_ISL_662424|20 ···················· ···················· ····················· ···················· ···················· ····················
T 6943 hCoV-19/England/QEUH-B67260/2020|EPI_ISL_662428|20 ···················· ···················· ····················· ···················· ···T················ ····················
T 6944 hCoV-19/England/ALDP-B733AA/2020|EPI_ISL_662429|20 ···················· ···················· ····················· ···················· ···················· ····················
T 6945 hCoV-19/England/QEUH-B6A625/2020|EPI_ISL_662430|20 ···················· ···················· ····················· ···················· ···················· ····················
T 6946 hCoV-19/England/ALDP-B73771/2020|EPI_ISL_662431|20 ···················· ···················· ····················· ···················· ···················· ····················
T 6947 hCoV-19/England/QEUH-B6B7B8/2020|EPI_ISL_662432|20 ···················· ···················· ····················· ···················· ···················· ····················
T 6948 hCoV-19/England/QEUH-B672E8/2020|EPI_ISL_662435|20 ···················· ···················· ····················· ···················· ···················· ····················
T 6949 hCoV-19/England/QEUH-B6B1AB/2020|EPI_ISL_662436|20 ···················· ···················· ····················· ···················· ···················· ····················
T 6950 hCoV-19/England/QEUH-B6A1D9/2020|EPI_ISL_662437|20 ···················· ···················· ····················· ···················· ···················· ····················
T 6951 hCoV-19/England/QEUH-B6A7C8/2020|EPI_ISL_662438|20 ···················· ···················· ····················· ···················· ···················· ····················
T 6952 hCoV-19/England/QEUH-B6729D/2020|EPI_ISL_662439|20 ···················· ···················· ····················· ···················· ···················· ····················
T 6953 hCoV-19/England/QEUH-B6A643/2020|EPI_ISL_662440|20 ···················· ·A·················· ····················· ···················· ···················· ····················
T 6954 hCoV-19/England/QEUH-B6AD8A/2020|EPI_ISL_662441|20 ···················· ···················· ····················· ···················· ···················· ····················
T 6955 hCoV-19/England/QEUH-B6A42B/2020|EPI_ISL_662442|20 ···················· ···················· ····················· ···················· ···················· ····················
T 6956 hCoV-19/England/QEUH-B6AD99/2020|EPI_ISL_662443|20 ···················· ···················· ····················· ···················· ···················· ····················
T 6957 hCoV-19/Australia/VIC17119/2020|EPI_ISL_663742|202 ···················· ···················· ····················· ···················· ···················· ····················
T 6958 hCoV-19/Australia/VIC17120/2020|EPI_ISL_663743|202 ···················· ···················· ····················· ···················· ···················· ····················
T 6959 hCoV-19/Australia/VIC17122/2020|EPI_ISL_663744|202 ···················· ···················· ····················· ···················· ···················· ····················
T 6960 hCoV-19/Australia/VIC17123/2020|EPI_ISL_663745|202 ···················· ···················· ····················· ···················· ···················· ····················
T 6961 hCoV-19/Australia/VIC17124/2020|EPI_ISL_663746|202 ···················· ···················· ····················· ···················· ···················· ····················
T 6962 hCoV-19/Australia/VIC17125/2020|EPI_ISL_663747|202 ···················· ···················· ····················· ···················· ···················· ····················
T 6963 hCoV-19/Australia/VIC17126/2020|EPI_ISL_663748|202 ·······Y············ ···················· ····················· ···················· ···················· ····················
T 6964 hCoV-19/Australia/VIC17127/2020|EPI_ISL_663749|202 ···················· ···················· ····················· ···················· ···················· ····················
T 6965 hCoV-19/Australia/VIC17128/2020|EPI_ISL_663750|202 ···················· ···················· ····················· ···················· ···················· ····················
T 6966 hCoV-19/Australia/VIC17130/2020|EPI_ISL_663751|202 ···················· ···················· ····················· ···················· ···················· ····················
T 6967 hCoV-19/Australia/VIC17131/2020|EPI_ISL_663752|202 ···················· ···················· ····················· ···················· ···················· ····················

| | | | | | | | | |
|---|---|---|---|---|---|---|---|---|
| T | 6968 | hCoV-19/Australia/VIC17132/2020\|EPI_ISL_663753\|202 | ···················· | ···················· | ···················· | ···················· | ···················· | ···················· |
| T | 6969 | hCoV-19/Australia/VIC17133/2020\|EPI_ISL_663754\|202 | ···················· | ···················· | ···················· | ···················· | ···················· | ···················· |
| T | 6970 | hCoV-19/Australia/VIC17134/2020\|EPI_ISL_663755\|202 | ···················· | ···················· | ···················· | ···················· | ···················· | ···················· |
| T | 6971 | hCoV-19/Australia/VIC17137/2020\|EPI_ISL_663756\|202 | ···················· | ···················· | ···················· | ···················· | ···················· | ···················· |
| T | 6972 | hCoV-19/Australia/VIC17138/2020\|EPI_ISL_663757\|202 | ···················· | ···················· | ···················· | ···················· | ···················· | ···················· |
| T | 6973 | hCoV-19/Australia/VIC17141/2020\|EPI_ISL_663758\|202 | ···················· | ···················· | ···················· | ···················· | ···················· | ···················· |
| T | 6974 | hCoV-19/Australia/VIC17142/2020\|EPI_ISL_663759\|202 | ···················· | ···················· | ···················· | ···················· | ···················· | ···················· |
| T | 6975 | hCoV-19/Australia/VIC17144/2020\|EPI_ISL_663760\|202 | ···················· | ···················· | ···················· | ···················· | ···················· | ···················· |
| T | 6976 | hCoV-19/Australia/VIC17145/2020\|EPI_ISL_663761\|202 | ···················· | ···················· | ···················· | ···················· | ···················· | ···················· |
| T | 6977 | hCoV-19/Australia/VIC17146/2020\|EPI_ISL_663762\|202 | ···················· | ···················· | ···················· | ···················· | ···················· | ···················· |
| T | 6978 | hCoV-19/Australia/VIC17147/2020\|EPI_ISL_663763\|202 | ···················· | ···················· | ···················· | ···················· | ···················· | ···················· |
| T | 6979 | hCoV-19/Australia/VIC17148/2020\|EPI_ISL_663764\|202 | ···················· | ···················· | ···················· | ···················· | ···················· | ···················· |
| T | 6980 | hCoV-19/Australia/VIC17150/2020\|EPI_ISL_663766\|202 | ···················· | ···················· | ···················· | ···················· | ···················· | ···················· |
| T | 6981 | hCoV-19/Australia/VIC17151/2020\|EPI_ISL_663767\|202 | ···················· | ···················· | ···················· | ···················· | ···················· | ···················· |
| T | 6982 | hCoV-19/Australia/VIC17153/2020\|EPI_ISL_663768\|202 | ···················· | ···················· | ···················· | ···················· | ···················· | ···················· |
| T | 6983 | hCoV-19/Australia/VIC17154/2020\|EPI_ISL_663769\|202 | ···················· | ···················· | ···················· | ···················· | ···················· | ···················· |
| T | 6984 | hCoV-19/Australia/VIC17156/2020\|EPI_ISL_663770\|202 | ···················· | ···················· | ···················· | ···················· | ···················· | ···················· |
| T | 6985 | hCoV-19/Australia/VIC17158/2020\|EPI_ISL_663771\|202 | ···················· | ···················· | ···················· | ···················· | ···················· | ···················· |
| T | 6986 | hCoV-19/Australia/VIC17159/2020\|EPI_ISL_663772\|202 | ···················· | ···················· | ···················· | ···················· | ···················· | ···················· |
| T | 6987 | hCoV-19/Australia/VIC17161/2020\|EPI_ISL_663773\|202 | ···················· | ···················· | ···················· | ···················· | ···················· | ···················· |
| T | 6988 | hCoV-19/Australia/VIC17162/2020\|EPI_ISL_663774\|202 | ···················· | ···················· | ···················· | ···················· | ···················· | ···················· |
| T | 6989 | hCoV-19/USA/OR-OHSU-2433/2020\|EPI_ISL_667207\|2020- | ···················· | ···················· | ···················· | ···················· | ···················· | ···················· |
| T | 6990 | hCoV-19/USA/OR-OHSU-2441/2020\|EPI_ISL_667208\|2020- | ···················· | ···················· | ···················· | ···················· | ···················· | ···················· |
| T | 6991 | hCoV-19/USA/OR-OHSU-2443/2020\|EPI_ISL_667209\|2020- | ···················· | ···················· | ···················· | ···················· | ···················· | ···················· |
| T | 6992 | hCoV-19/USA/OR-OHSU-2445/2020\|EPI_ISL_667210\|2020- | ···················· | ···················· | ···················· | ···················· | ···················· | ···················· |
| T | 6993 | hCoV-19/USA/OR-OHSU-2451/2020\|EPI_ISL_667212\|2020- | ···················· | ···················· | ···················· | ···················· | ···················· | ···················· |
| T | 6994 | hCoV-19/USA/OR-OHSU-2457/2020\|EPI_ISL_667213\|2020- | ···················· | ·A·················· | ···················· | ···················· | ···················· | ···················· |
| T | 6995 | hCoV-19/USA/OR-OHSU-2463/2020\|EPI_ISL_667216\|2020- | ···················· | ·A·················· | ···················· | ···················· | ···················· | ···················· |
| T | 6996 | hCoV-19/USA/OR-OHSU-2465/2020\|EPI_ISL_667217\|2020- | ···················· | ···················· | ···················· | ···················· | ···················· | ···················· |
| T | 6997 | hCoV-19/USA/OR-OHSU-2467/2020\|EPI_ISL_667218\|2020- | ···················· | ·A·················· | ···················· | ···················· | ···················· | ···················· |
| T | 6998 | hCoV-19/USA/OR-OHSU-2471/2020\|EPI_ISL_667219\|2020- | ···················· | ···················· | ···················· | ···················· | ···················· | ···················· |
| T | 6999 | hCoV-19/USA/OR-OHSU-2473/2020\|EPI_ISL_667220\|2020- | ···················· | ·A·················· | ···················· | ···················· | ···················· | ···················· |
| T | 7000 | hCoV-19/USA/WA-OHSU-2479/2020\|EPI_ISL_667222\|2020- | ···················· | ·A·················· | ···················· | ···················· | ···················· | ···················· |
| T | 7001 | hCoV-19/USA/OR-OHSU-2481/2020\|EPI_ISL_667223\|2020- | ···················· | ···················· | ···················· | ···················· | ···················· | ···················· |
| T | 7002 | hCoV-19/USA/OR-OHSU-2483/2020\|EPI_ISL_667224\|2020- | ···················· | ·A·················· | ···················· | ···················· | ···················· | ···················· |
| T | 7003 | hCoV-19/USA/OR-OHSU-2487/2020\|EPI_ISL_667225\|2020- | ···················· | ·A·················· | ···················· | ···················· | ···················· | ···················· |
| T | 7004 | hCoV-19/USA/OR-OHSU-2489/2020\|EPI_ISL_667226\|2020- | ···················· | ···················· | ···················· | ···················· | ···················· | ···················· |
| T | 7005 | hCoV-19/USA/WA-OHSU-2501/2020\|EPI_ISL_667229\|2020- | ···················· | ·A·················· | ···················· | ···················· | ···················· | ···················· |
| T | 7006 | hCoV-19/USA/OR-OHSU-2503/2020\|EPI_ISL_667230\|2020- | ···················· | ···················· | ···················· | ···················· | ···················· | ···················· |
| T | 7007 | hCoV-19/USA/OR-OHSU-2504/2020\|EPI_ISL_667231\|2020- | ···················· | ···················· | ···················· | ···················· | ···················· | ···················· |
| T | 7008 | hCoV-19/USA/OR-OHSU-2507/2020\|EPI_ISL_667232\|2020- | ···················· | ·A·················· | ···················· | ···················· | ···················· | ···················· |
| T | 7009 | hCoV-19/USA/OR-OHSU-2511/2020\|EPI_ISL_667233\|2020- | ···················· | ·A·················· | ···················· | ···················· | ···················· | ···················· |
| T | 7010 | hCoV-19/USA/OR-OHSU-2513/2020\|EPI_ISL_667234\|2020- | ···················· | ···················· | ···················· | ···················· | ···················· | ···················· |
| T | 7011 | hCoV-19/USA/OR-OHSU-2517/2020\|EPI_ISL_667235\|2020- | ···················· | ···················· | ···················· | ···················· | ···················· | ···················· |
| T | 7012 | hCoV-19/USA/OR-OHSU-2523/2020\|EPI_ISL_667237\|2020- | ···················· | ···················· | ···················· | ···················· | ···················· | ···················· |
| T | 7013 | hCoV-19/USA/OR-OHSU-2527/2020\|EPI_ISL_667239\|2020- | ···················· | ·A·················· | ···················· | ···················· | ···················· | ···················· |
| T | 7014 | hCoV-19/USA/OR-OHSU-2529/2020\|EPI_ISL_667240\|2020- | ···················· | ···················· | ···················· | ···················· | ···················· | ···················· |
| T | 7015 | hCoV-19/USA/OR-OHSU-2535/2020\|EPI_ISL_667241\|2020- | ···················· | ·A·················· | ···················· | ···················· | ···················· | ···················· |
| T | 7016 | hCoV-19/USA/OR-OHSU-2537/2020\|EPI_ISL_667242\|2020- | ···················· | ···················· | ···················· | ···················· | ···················· | ···················· |
| T | 7017 | hCoV-19/USA/OR-OHSU-2539/2020\|EPI_ISL_667243\|2020- | ···················· | ···················· | ···················· | ···················· | ···················· | ···················· |
| T | 7018 | hCoV-19/USA/OR-OHSU-2541/2020\|EPI_ISL_667244\|2020- | ···················· | ···················· | ···················· | ···················· | ···················· | ···················· |
| T | 7019 | hCoV-19/USA/OR-OHSU-2543/2020\|EPI_ISL_667245\|2020- | ···················· | ···················· | ···················· | ···················· | ···················· | ···················· |
| T | 7020 | hCoV-19/USA/OR-OHSU-2549/2020\|EPI_ISL_667247\|2020- | ···················· | ···················· | ···················· | ···················· | ···················· | ···················· |
| T | 7021 | hCoV-19/USA/OR-OHSU-2551/2020\|EPI_ISL_667248\|2020- | ···················· | ···················· | ···················· | ···················· | ···················· | ···················· |
| T | 7022 | hCoV-19/USA/OR-OHSU-2557/2020\|EPI_ISL_667251\|2020- | ···················· | ···················· | ···················· | ···················· | ···················· | ···················· |
| T | 7023 | hCoV-19/USA/OR-OHSU-2559/2020\|EPI_ISL_667252\|2020- | ···················· | ·A·················· | ···················· | ···················· | ···················· | ···················· |
| T | 7024 | hCoV-19/USA/OR-OHSU-2563/2020\|EPI_ISL_667253\|2020- | ···················· | ···················· | ···················· | ···················· | ···················· | ···················· |
| T | 7025 | hCoV-19/USA/OR-OHSU-2569/2020\|EPI_ISL_667254\|2020- | ···················· | ···················· | ···················· | ···················· | ···················· | ···················· |
| T | 7026 | hCoV-19/USA/OR-OHSU-2575/2020\|EPI_ISL_667255\|2020- | ···················· | ···················· | ···················· | ···················· | ···················· | ···················· |
| T | 7027 | hCoV-19/USA/OR-OHSU-2577/2020\|EPI_ISL_667256\|2020- | ···················· | ·A·················· | ···················· | ···················· | ···················· | ···················· |
| T | 7028 | hCoV-19/USA/OR-OHSU-2579/2020\|EPI_ISL_667257\|2020- | ···················· | ···················· | ···················· | ···················· | ···················· | ···················· |
| T | 7029 | hCoV-19/USA/OR-OHSU-2589/2020\|EPI_ISL_667258\|2020- | ···················· | ···················· | ···················· | ···················· | ···················· | ···················· |
| T | 7030 | hCoV-19/USA/OR-OHSU-2597/2020\|EPI_ISL_667260\|2020- | ···················· | ···················· | ···················· | ···················· | ···················· | ···················· |
| T | 7031 | hCoV-19/USA/OR-OHSU-2599/2020\|EPI_ISL_667261\|2020- | ···················· | ···················· | ···················· | ···················· | ···················· | ···················· |
| T | 7032 | hCoV-19/USA/WA-OHSU-2603/2020\|EPI_ISL_667262\|2020- | ···················· | ···················· | ···················· | ···················· | ···················· | ···················· |
| T | 7033 | hCoV-19/USA/OR-OHSU-2607/2020\|EPI_ISL_667263\|2020- | ···················· | ·A·················· | ···················· | ···················· | ···················· | ···················· |
| T | 7034 | hCoV-19/USA/OR-OHSU-2619/2020\|EPI_ISL_667264\|2020- | ···················· | ·A·················· | ···················· | ···················· | ···················· | ···················· |
| T | 7035 | hCoV-19/USA/OR-OHSU-2621/2020\|EPI_ISL_667265\|2020- | ···················· | ·A·················· | ···················· | ···················· | ···················· | ···················· |
| T | 7036 | hCoV-19/USA/OR-OHSU-2623/2020\|EPI_ISL_667266\|2020- | ···················· | ·A·················· | ···················· | ···················· | ···················· | ···················· |
| T | 7037 | hCoV-19/USA/OR-OHSU-2625/2020\|EPI_ISL_667267\|2020- | ···················· | ···················· | ···················· | ···················· | ···················· | ···················· |
| T | 7038 | hCoV-19/USA/OR-OHSU-2631/2020\|EPI_ISL_667268\|2020- | ···················· | ·A·················· | ···················· | ···················· | ···················· | ···················· |
| T | 7039 | hCoV-19/USA/OR-OHSU-2637/2020\|EPI_ISL_667269\|2020- | ···················· | ···················· | ···················· | ···················· | ···················· | ···················· |
| T | 7040 | hCoV-19/Denmark/DCGC-10249/2020\|EPI_ISL_669817\|202 | ···················· | ·A·················· | ···················· | ···················· | ···················· | ···················· |
| T | 7041 | hCoV-19/Denmark/DCGC-10212/2020\|EPI_ISL_669834\|202 | ···················· | ···················· | ···················· | ···················· | ···················· | ···················· |
| T | 7042 | hCoV-19/Denmark/DCGC-10214/2020\|EPI_ISL_669835\|202 | ···················· | ···················· | ···················· | ···················· | ···················· | ···················· |
| T | 7043 | hCoV-19/Denmark/DCGC-10216/2020\|EPI_ISL_669836\|202 | ···················· | ···················· | ···················· | ···················· | ···················· | ···················· |
| T | 7044 | hCoV-19/Denmark/DCGC-10217/2020\|EPI_ISL_669837\|202 | ···················· | ···················· | ···················· | ···················· | ···················· | ···················· |
| T | 7045 | hCoV-19/Denmark/DCGC-10218/2020\|EPI_ISL_669838\|202 | ···················· | ···················· | ···················· | ···················· | ···················· | ···················· |
| T | 7046 | hCoV-19/Denmark/DCGC-10220/2020\|EPI_ISL_669839\|202 | ···················· | ···················· | ···················· | ···················· | ···················· | ···················· |
| T | 7047 | hCoV-19/Denmark/DCGC-10221/2020\|EPI_ISL_669840\|202 | ···················· | ···················· | ···················· | ···················· | ···················· | ···················· |

| | | | | | | | |
|---|---|---|---|---|---|---|---|
| T | 7048 hCoV-19/Denmark/DCGC-10222/2020\|EPI_ISL_669841\|202 | ···················· | ·A·················· | ···················· | ···················· | ···················· | ··A················· |
| T | 7049 hCoV-19/Denmark/DCGC-10224/2020\|EPI_ISL_669842\|202 | ···················· | ···················· | ···················· | ···················· | ···················· | ···················· |
| T | 7050 hCoV-19/Denmark/DCGC-10226/2020\|EPI_ISL_669843\|202 | ···················· | ···················· | ···················· | ···················· | ···················· | ···················· |
| T | 7051 hCoV-19/Denmark/DCGC-10227/2020\|EPI_ISL_669844\|202 | ···················· | ···················· | ···················· | ···················· | ···················· | ···················· |
| T | 7052 hCoV-19/Denmark/DCGC-10228/2020\|EPI_ISL_669845\|202 | ···················· | ···················· | ···················· | ···················· | ···················· | ···················· |
| T | 7053 hCoV-19/Denmark/DCGC-10229/2020\|EPI_ISL_669846\|202 | ···················· | ···················· | ···················· | ···················· | ···················· | ···················· |
| T | 7054 hCoV-19/Denmark/DCGC-10252/2020\|EPI_ISL_669858\|202 | ···················· | ···················· | ···················· | ···················· | ···················· | ···················· |
| T | 7055 hCoV-19/Denmark/DCGC-10253/2020\|EPI_ISL_669859\|202 | ···················· | ···················· | ···················· | ···················· | ···················· | ···················· |
| T | 7056 hCoV-19/Denmark/DCGC-10272/2020\|EPI_ISL_669869\|202 | ···················· | ···················· | ···················· | ···················· | ···················· | ···················· |
| T | 7057 hCoV-19/Denmark/DCGC-10273/2020\|EPI_ISL_669870\|202 | ···················· | ···················· | ···················· | ···················· | ···················· | ···················· |
| T | 7058 hCoV-19/Denmark/DCGC-10274/2020\|EPI_ISL_669871\|202 | ···················· | ···················· | ···················· | ···················· | ···················· | ···················· |
| T | 7059 hCoV-19/Denmark/DCGC-10278/2020\|EPI_ISL_669872\|202 | ···················· | ···················· | ···················· | ···················· | ···················· | ···················· |
| T | 7060 hCoV-19/Denmark/DCGC-10279/2020\|EPI_ISL_669873\|202 | ···················· | ···················· | ···················· | ···················· | ···················· | ···················· |
| T | 7061 hCoV-19/Denmark/DCGC-10281/2020\|EPI_ISL_669874\|202 | ···················· | ···················· | ···················· | ···················· | ···················· | ···················· |
| T | 7062 hCoV-19/Denmark/DCGC-10283/2020\|EPI_ISL_669875\|202 | ···················· | ···················· | ···················· | ···················· | ···················· | ···················· |
| T | 7063 hCoV-19/Denmark/DCGC-10284/2020\|EPI_ISL_669876\|202 | ···················· | ···················· | ···················· | ···················· | ···················· | ···················· |
| T | 7064 hCoV-19/Denmark/DCGC-10285/2020\|EPI_ISL_669877\|202 | ···················· | ···················· | ···················· | ···················· | ···················· | ···················· |
| T | 7065 hCoV-19/Denmark/DCGC-10287/2020\|EPI_ISL_669878\|202 | ···················· | ···················· | ···················· | ···················· | ···················· | ···················· |
| T | 7066 hCoV-19/Denmark/DCGC-10292/2020\|EPI_ISL_669879\|202 | ···················· | ···················· | ···················· | ···················· | ···················· | ···················· |
| T | 7067 hCoV-19/Denmark/DCGC-10293/2020\|EPI_ISL_669880\|202 | ···················· | ···················· | ···················· | ···················· | ···················· | ···················· |
| T | 7068 hCoV-19/Denmark/DCGC-10295/2020\|EPI_ISL_669881\|202 | ···················· | ···················· | ···················· | ···················· | ···················· | ···················· |
| T | 7069 hCoV-19/Denmark/DCGC-10296/2020\|EPI_ISL_669882\|202 | ···················· | ···················· | ···················· | ···················· | ···················· | ···················· |
| T | 7070 hCoV-19/Denmark/DCGC-10298/2020\|EPI_ISL_669883\|202 | ···················· | ···················· | ···················· | ···················· | ···················· | ···················· |
| T | 7071 hCoV-19/Denmark/DCGC-10299/2020\|EPI_ISL_669884\|202 | ···················· | ···················· | ···················· | ···················· | ···················· | ···················· |
| T | 7072 hCoV-19/Denmark/DCGC-10300/2020\|EPI_ISL_669885\|202 | ···················· | ···················· | ···················· | ···················· | ···················· | ···················· |
| T | 7073 hCoV-19/Denmark/DCGC-10301/2020\|EPI_ISL_669886\|202 | ···················· | ···················· | ···················· | ···················· | ···················· | ···················· |
| T | 7074 hCoV-19/Denmark/DCGC-10303/2020\|EPI_ISL_669887\|202 | ···················· | ···················· | ···················· | ···················· | ···················· | ···················· |
| T | 7075 hCoV-19/Denmark/DCGC-10305/2020\|EPI_ISL_669888\|202 | ···················· | ···················· | ···················· | ···················· | ···················· | ···················· |
| T | 7076 hCoV-19/Denmark/DCGC-10308/2020\|EPI_ISL_669889\|202 | ···················· | ···················· | ···················· | ···················· | ···················· | ···················· |
| T | 7077 hCoV-19/Denmark/DCGC-10309/2020\|EPI_ISL_669890\|202 | ···················· | ···················· | ···················· | ···················· | ···················· | ···················· |
| T | 7078 hCoV-19/Denmark/DCGC-10310/2020\|EPI_ISL_669891\|202 | ···················· | ···················· | ···················· | ···················· | ···················· | ···················· |
| T | 7079 hCoV-19/Denmark/DCGC-10311/2020\|EPI_ISL_669892\|202 | ···················· | ···················· | ···················· | ···················· | ···················· | ···················· |
| T | 7080 hCoV-19/Denmark/DCGC-10312/2020\|EPI_ISL_669893\|202 | ···················· | ···················· | ···················· | ···················· | ···················· | ···················· |
| T | 7081 hCoV-19/Denmark/DCGC-10313/2020\|EPI_ISL_669894\|202 | ···················· | ···················· | ···················· | ···················· | ···················· | ···················· |
| T | 7082 hCoV-19/Denmark/DCGC-10314/2020\|EPI_ISL_669895\|202 | ···················· | ···················· | ···················· | ···················· | ···················· | ···················· |
| T | 7083 hCoV-19/Denmark/DCGC-10316/2020\|EPI_ISL_669896\|202 | ···················· | ···················· | ···················· | ···················· | ···················· | ···················· |
| T | 7084 hCoV-19/Denmark/DCGC-10318/2020\|EPI_ISL_669897\|202 | ···················· | ···················· | ···················· | ···················· | ···················· | ···················· |
| T | 7085 hCoV-19/Denmark/DCGC-10319/2020\|EPI_ISL_669898\|202 | ···················· | ···················· | ···················· | ···················· | ···················· | ···················· |
| T | 7086 hCoV-19/Denmark/DCGC-10320/2020\|EPI_ISL_669899\|202 | ···················· | ···················· | ···················· | ···················· | ···················· | ···················· |
| T | 7087 hCoV-19/Denmark/DCGC-11261/2020\|EPI_ISL_670800\|202 | ···················· | ···················· | ···················· | ···················· | ···················· | ···················· |
| T | 7088 hCoV-19/Denmark/DCGC-11262/2020\|EPI_ISL_670801\|202 | ···················· | ·A·················· | ···················· | ···················· | ···················· | ···················· |
| T | 7089 hCoV-19/Denmark/DCGC-11265/2020\|EPI_ISL_670802\|202 | ···················· | ·A·················· | ···················· | ···················· | ···················· | ···················· |
| T | 7090 hCoV-19/Denmark/DCGC-11268/2020\|EPI_ISL_670803\|202 | ···················· | ···················· | ···················· | ···················· | ···················· | ···················· |
| T | 7091 hCoV-19/Denmark/DCGC-11269/2020\|EPI_ISL_670804\|202 | ···················· | ·A·················· | ···················· | ···················· | ···················· | ··A················· |
| T | 7092 hCoV-19/Denmark/DCGC-11272/2020\|EPI_ISL_670805\|202 | ···················· | ···················· | ···················· | ···················· | ···················· | ···················· |
| T | 7093 hCoV-19/Denmark/DCGC-11273/2020\|EPI_ISL_670806\|202 | ···················· | ·A·················· | ···················· | ···················· | ···················· | ···················· |
| T | 7094 hCoV-19/Denmark/DCGC-11274/2020\|EPI_ISL_670807\|202 | ···················· | ···················· | ···················· | ···················· | ···················· | ···················· |
| T | 7095 hCoV-19/Denmark/DCGC-11275/2020\|EPI_ISL_670808\|202 | ···················· | ···················· | ···················· | ···················· | ···················· | ···················· |
| T | 7096 hCoV-19/Denmark/DCGC-11278/2020\|EPI_ISL_670809\|202 | ···················· | ···················· | ···················· | ···················· | ···················· | ···················· |
| T | 7097 hCoV-19/Denmark/DCGC-11280/2020\|EPI_ISL_670810\|202 | ···················· | ···················· | ···················· | ···················· | ···················· | ···················· |
| T | 7098 hCoV-19/Denmark/DCGC-11282/2020\|EPI_ISL_670811\|202 | ···················· | ···················· | ···················· | ···················· | ···················· | ···················· |
| T | 7099 hCoV-19/Denmark/DCGC-11283/2020\|EPI_ISL_670812\|202 | ···················· | ···················· | ···················· | ···················· | ···················· | ···················· |
| T | 7100 hCoV-19/Denmark/DCGC-11286/2020\|EPI_ISL_670813\|202 | ···················· | ···················· | ···················· | ···················· | ···················· | ···················· |
| T | 7101 hCoV-19/Denmark/DCGC-11288/2020\|EPI_ISL_670814\|202 | ···················· | ···················· | ···················· | ···················· | ···················· | ···················· |
| T | 7102 hCoV-19/Denmark/DCGC-11289/2020\|EPI_ISL_670815\|202 | ···················· | ···················· | ···················· | ···················· | ···················· | ···················· |
| T | 7103 hCoV-19/Denmark/DCGC-11290/2020\|EPI_ISL_670816\|202 | ···················· | ···················· | ···················· | ···················· | ···················· | ···················· |
| T | 7104 hCoV-19/Denmark/DCGC-11291/2020\|EPI_ISL_670817\|202 | ···················· | ···················· | ···················· | ···················· | ···················· | ···················· |
| T | 7105 hCoV-19/Denmark/DCGC-11292/2020\|EPI_ISL_670818\|202 | ···················· | ···················· | ···················· | ···················· | ···················· | ···················· |
| T | 7106 hCoV-19/Denmark/DCGC-11293/2020\|EPI_ISL_670819\|202 | ···················· | ···················· | ···················· | ···················· | ···················· | ···················· |
| T | 7107 hCoV-19/Denmark/DCGC-11295/2020\|EPI_ISL_670820\|202 | ···················· | ···················· | ···················· | ···················· | ···················· | ···················· |
| T | 7108 hCoV-19/Denmark/DCGC-11296/2020\|EPI_ISL_670821\|202 | ···················· | ···················· | ···················· | ···················· | ···················· | ···················· |
| T | 7109 hCoV-19/Denmark/DCGC-11297/2020\|EPI_ISL_670822\|202 | ···················· | ·A·················· | ···················· | ···················· | ···················· | ···················· |
| T | 7110 hCoV-19/Denmark/DCGC-11299/2020\|EPI_ISL_670823\|202 | ···················· | ······C············· | ···················· | ···················· | ···················· | ···················· |
| T | 7111 hCoV-19/Denmark/DCGC-11300/2020\|EPI_ISL_670824\|202 | ···················· | ···················· | ···················· | ···················· | ···················· | ···················· |
| T | 7112 hCoV-19/Denmark/DCGC-11301/2020\|EPI_ISL_670825\|202 | ···················· | ···················· | ···················· | ···················· | ···················· | ···················· |
| T | 7113 hCoV-19/Denmark/DCGC-11302/2020\|EPI_ISL_670826\|202 | ···················· | ·A·················· | ···················· | ···················· | ···················· | ··A················· |
| T | 7114 hCoV-19/Denmark/DCGC-11305/2020\|EPI_ISL_670827\|202 | ···················· | ···················· | ···················· | ···················· | ···················· | ···················· |
| T | 7115 hCoV-19/Denmark/DCGC-11306/2020\|EPI_ISL_670828\|202 | ···················· | ···················· | ···················· | ···················· | ···················· | ···················· |
| T | 7116 hCoV-19/Denmark/DCGC-11307/2020\|EPI_ISL_670829\|202 | ···················· | ···················· | ···················· | ···················· | ···················· | ···················· |
| T | 7117 hCoV-19/Denmark/DCGC-11334/2020\|EPI_ISL_670846\|202 | ···················· | ···················· | ···················· | ···················· | ···················· | ···················· |
| T | 7118 hCoV-19/Denmark/DCGC-11340/2020\|EPI_ISL_670849\|202 | ···················· | ···················· | ···················· | ···················· | ···················· | ···················· |
| T | 7119 hCoV-19/Denmark/DCGC-11341/2020\|EPI_ISL_670850\|202 | ···················· | ···················· | ···················· | ···················· | ···················· | ···················· |
| T | 7120 hCoV-19/Denmark/DCGC-11342/2020\|EPI_ISL_670851\|202 | ···················· | ···················· | ···················· | ···················· | ···················· | ···················· |
| T | 7121 hCoV-19/Denmark/DCGC-11343/2020\|EPI_ISL_670852\|202 | ···················· | ···················· | ···················· | ···················· | ···················· | ···················· |
| T | 7122 hCoV-19/Denmark/DCGC-11344/2020\|EPI_ISL_670853\|202 | ···················· | ···················· | ···················· | ···················· | ···················· | ···················· |
| T | 7123 hCoV-19/Denmark/DCGC-11345/2020\|EPI_ISL_670854\|202 | ···················· | ···················· | ···················· | ···················· | ···················· | ···················· |
| T | 7124 hCoV-19/Denmark/DCGC-11347/2020\|EPI_ISL_670855\|202 | ···················· | ···················· | ···················· | ···················· | ···················· | ···················· |
| T | 7125 hCoV-19/Denmark/DCGC-11348/2020\|EPI_ISL_670856\|202 | ···················· | ···················· | ···················· | ···················· | ···················· | ···················· |
| T | 7126 hCoV-19/Denmark/DCGC-11349/2020\|EPI_ISL_670857\|202 | ···················· | ·A·················· | ···················· | ···················· | ···················· | ···················· |
| T | 7127 hCoV-19/Denmark/DCGC-11350/2020\|EPI_ISL_670858\|202 | ···················· | ···················· | ···················· | ···················· | ···················· | ···················· |

| | | | | | | | | | |
|---|---|---|---|---|---|---|---|---|---|
| T | 7128 | hCoV-19/Denmark/DCGC-11351/2020\|EPI_ISL_670859\|202 | ···················· | ·A·················· | ····················· | ···················· | ···················· | ···················· | |
| T | 7129 | hCoV-19/Denmark/DCGC-11353/2020\|EPI_ISL_670860\|202 | ···················· | ···················· | ····················· | ···················· | ···················· | ···················· | |
| T | 7130 | hCoV-19/Denmark/DCGC-11354/2020\|EPI_ISL_670861\|202 | ···················· | ···················· | ····················· | ···················· | ···················· | ···················· | |
| T | 7131 | hCoV-19/Denmark/DCGC-11355/2020\|EPI_ISL_670862\|202 | ···················· | ···················· | ····················· | ···················· | ···················· | ···················· | |
| T | 7132 | hCoV-19/Denmark/DCGC-11357/2020\|EPI_ISL_670863\|202 | ···················· | ···················· | ····················· | ···················· | ···················· | ···················· | |
| T | 7133 | hCoV-19/Denmark/DCGC-11358/2020\|EPI_ISL_670864\|202 | ···················· | ···················· | ····················· | ···················· | ···················· | ···················· | |
| T | 7134 | hCoV-19/Denmark/DCGC-11359/2020\|EPI_ISL_670865\|202 | ···················· | ···················· | ····················· | ···················· | ···················· | ···················· | |
| T | 7135 | hCoV-19/Denmark/DCGC-11361/2020\|EPI_ISL_670866\|202 | ···················· | ···················· | ····················· | ···················· | ···················· | ···················· | |
| T | 7136 | hCoV-19/Denmark/DCGC-11362/2020\|EPI_ISL_670867\|202 | ···················· | ···················· | ····················· | ···················· | ···················· | ···················· | |
| T | 7137 | hCoV-19/Denmark/DCGC-11364/2020\|EPI_ISL_670868\|202 | ···················· | ···················· | ····················· | ···················· | ···················· | ···················· | |
| T | 7138 | hCoV-19/Denmark/DCGC-11365/2020\|EPI_ISL_670869\|202 | ···················· | ···················· | ····················· | ···················· | ···················· | ···················· | |
| T | 7139 | hCoV-19/Denmark/DCGC-11370/2020\|EPI_ISL_670870\|202 | ···················· | ···················· | ····················· | ···················· | ···················· | ···················· | |
| T | 7140 | hCoV-19/Denmark/DCGC-11372/2020\|EPI_ISL_670871\|202 | ···················· | ···················· | ····················· | ···················· | ···················· | ···················· | |
| T | 7141 | hCoV-19/Denmark/DCGC-11373/2020\|EPI_ISL_670872\|202 | ···················· | ···················· | ····················· | ···················· | ···················· | ···················· | |
| T | 7142 | hCoV-19/Denmark/DCGC-11374/2020\|EPI_ISL_670873\|202 | ···················· | ···················· | ····················· | ···················· | ···················· | ···················· | |
| T | 7143 | hCoV-19/Denmark/DCGC-11376/2020\|EPI_ISL_670874\|202 | ···················· | ···················· | ····················· | ···················· | ···················· | ···················· | |
| T | 7144 | hCoV-19/Denmark/DCGC-11377/2020\|EPI_ISL_670875\|202 | ···················· | ···················· | ····················· | ···················· | ···················· | ···················· | |
| T | 7145 | hCoV-19/Denmark/DCGC-11378/2020\|EPI_ISL_670876\|202 | ···················· | ···················· | ····················· | ···················· | ···················· | ···················· | |
| T | 7146 | hCoV-19/Denmark/DCGC-11379/2020\|EPI_ISL_670877\|202 | ···················· | ···················· | ····················· | ···················· | ···················· | ···················· | |
| T | 7147 | hCoV-19/Denmark/DCGC-11381/2020\|EPI_ISL_670878\|202 | ···················· | ···················· | ····················· | ···················· | ···················· | ···················· | |
| T | 7148 | hCoV-19/Denmark/DCGC-11382/2020\|EPI_ISL_670879\|202 | ···················· | ···················· | ····················· | ···················· | ···················· | ···················· | |
| T | 7149 | hCoV-19/Denmark/DCGC-11384/2020\|EPI_ISL_670880\|202 | ···················· | ···················· | ····················· | ···················· | ···················· | ···················· | |
| T | 7150 | hCoV-19/Denmark/DCGC-11385/2020\|EPI_ISL_670881\|202 | ···················· | ···················· | ····················· | ···················· | ···················· | ···················· | |
| T | 7151 | hCoV-19/Denmark/DCGC-11387/2020\|EPI_ISL_670882\|202 | ···················· | ···················· | ····················· | ···················· | ···················· | ···················· | |
| T | 7152 | hCoV-19/Denmark/DCGC-11389/2020\|EPI_ISL_670883\|202 | ···················· | ···················· | ····················· | ···················· | ···················· | ···················· | |
| T | 7153 | hCoV-19/Denmark/DCGC-11390/2020\|EPI_ISL_670884\|202 | ···················· | ···················· | ····················· | ···················· | ···················· | ···················· | |
| T | 7154 | hCoV-19/Denmark/DCGC-11394/2020\|EPI_ISL_670885\|202 | ···················· | ···················· | ····················· | ···················· | ···················· | ···················· | |
| T | 7155 | hCoV-19/Denmark/DCGC-11395/2020\|EPI_ISL_670886\|202 | ···················· | ···················· | ····················· | ···················· | ···················· | ···················· | |
| T | 7156 | hCoV-19/Denmark/DCGC-11397/2020\|EPI_ISL_670887\|202 | ···················· | ···················· | ····················· | ···················· | ···················· | ···················· | |
| T | 7157 | hCoV-19/Denmark/DCGC-11398/2020\|EPI_ISL_670888\|202 | ···················· | ···················· | ····················· | ···················· | ···················· | ···················· | |
| T | 7158 | hCoV-19/Denmark/DCGC-11399/2020\|EPI_ISL_670889\|202 | ···················· | ···················· | ····················· | ···················· | ···················· | ···················· | |
| T | 7159 | hCoV-19/Denmark/DCGC-11400/2020\|EPI_ISL_670890\|202 | ···················· | ···················· | ····················· | ···················· | ···················· | ···················· | |
| T | 7160 | hCoV-19/Denmark/DCGC-11402/2020\|EPI_ISL_670891\|202 | ···················· | ···················· | ····················· | ···················· | ···················· | ···················· | |
| T | 7161 | hCoV-19/Denmark/DCGC-11403/2020\|EPI_ISL_670892\|202 | ···················· | ···················· | ····················· | ···················· | ···················· | ···················· | |
| T | 7162 | hCoV-19/Denmark/DCGC-11404/2020\|EPI_ISL_670893\|202 | ···················· | ·A·················· | ····················· | ···················· | ···················· | ···················· | |
| T | 7163 | hCoV-19/Denmark/DCGC-11405/2020\|EPI_ISL_670894\|202 | ···················· | ···················· | ····················· | ···················· | ···················· | ···················· | |
| T | 7164 | hCoV-19/Denmark/DCGC-11406/2020\|EPI_ISL_670895\|202 | ···················· | ·A·················· | ····················· | ···················· | ···················· | ···················· | |
| T | 7165 | hCoV-19/Denmark/DCGC-11407/2020\|EPI_ISL_670896\|202 | ···················· | ···················· | ····················· | ···················· | ···················· | ···················· | |
| T | 7166 | hCoV-19/Denmark/DCGC-11408/2020\|EPI_ISL_670897\|202 | ···················· | ···················· | ····················· | ···················· | ···················· | ···················· | |
| T | 7167 | hCoV-19/Denmark/DCGC-11409/2020\|EPI_ISL_670898\|202 | ···················· | ···················· | ····················· | ···················· | ···················· | ···················· | |
| T | 7168 | hCoV-19/Denmark/DCGC-11410/2020\|EPI_ISL_670899\|202 | ···················· | ···················· | ····················· | ···················· | ···················· | ···················· | |
| T | 7169 | hCoV-19/England/QEUH-B66B57/2020\|EPI_ISL_662323\|20 | ···················· | ···················· | ····················· | ···················· | ···················· | ···················· | |
| T | 7170 | hCoV-19/England/QEUH-B6A9B3/2020\|EPI_ISL_662324\|20 | ···················· | ···················· | ····················· | ···················· | ···················· | ···················· | |
| T | 7171 | hCoV-19/England/QEUH-B6AE0F/2020\|EPI_ISL_662325\|20 | ···················· | ···················· | ····················· | ···················· | ···················· | ···················· | |
| T | 7172 | hCoV-19/England/QEUH-B66BA2/2020\|EPI_ISL_662326\|20 | ···················· | ···················· | ····················· | ···················· | ···················· | ···················· | |
| T | 7173 | hCoV-19/England/ALDP-B7347A/2020\|EPI_ISL_662327\|20 | ···················· | ···················· | ····················· | ···················· | ····T··············· | ···················· | |
| T | 7174 | hCoV-19/England/ALDP-B736A1/2020\|EPI_ISL_662334\|20 | ···················· | ···················· | ····················· | ···················· | ···················· | ···················· | |
| T | 7175 | hCoV-19/England/QEUH-B66850/2020\|EPI_ISL_662335\|20 | ···················· | ·A·················· | ····················· | ···················· | ···················· | ···A················ | |
| T | 7176 | hCoV-19/England/QEUH-B6735E/2020\|EPI_ISL_662337\|20 | ···················· | ·A·················· | ····················· | ···················· | ···················· | ···A················ | |
| T | 7177 | hCoV-19/England/QEUH-B6AFC0/2020\|EPI_ISL_662338\|20 | ···················· | ···················· | ····················· | ···················· | ···················· | ···················· | |
| T | 7178 | hCoV-19/England/QEUH-B67594/2020\|EPI_ISL_662339\|20 | ···················· | ···················· | ····················· | ···················· | ···················· | ···················· | |
| T | 7179 | hCoV-19/England/QEUH-B6AFDF/2020\|EPI_ISL_662340\|20 | ···················· | ···················· | ····················· | ···················· | ···················· | ···················· | |
| T | 7180 | hCoV-19/England/QEUH-B67628/2020\|EPI_ISL_662341\|20 | ···················· | ···················· | ····················· | ···················· | ···················· | ···················· | |
| T | 7181 | hCoV-19/England/ALDP-B73410/2020\|EPI_ISL_662342\|20 | ···················· | ···················· | ····················· | ···················· | ···················· | ···················· | |
| T | 7182 | hCoV-19/England/ALDP-B73708/2020\|EPI_ISL_662343\|20 | ···················· | ···················· | ····················· | ···················· | ···················· | ···················· | |
| T | 7183 | hCoV-19/England/ALDP-B73A5A/2020\|EPI_ISL_662344\|20 | ···················· | ···················· | ····················· | ···················· | ···················· | ···················· | |
| T | 7184 | hCoV-19/England/QEUH-B6A977/2020\|EPI_ISL_662345\|20 | ···················· | ···················· | ····················· | ···················· | ···················· | ···················· | |
| T | 7185 | hCoV-19/England/QEUH-B6A212/2020\|EPI_ISL_662347\|20 | ···················· | ···················· | ····················· | ···················· | ···················· | ···················· | |
| T | 7186 | hCoV-19/England/QEUH-B6687E/2020\|EPI_ISL_662348\|20 | ···················· | ···················· | ····················· | ···················· | ···················· | ···················· | |
| T | 7187 | hCoV-19/England/QEUH-B67321/2020\|EPI_ISL_662349\|20 | ···················· | ···················· | ····················· | ···················· | ···················· | ···················· | |
| T | 7188 | hCoV-19/England/QEUH-B6A564/2020\|EPI_ISL_662350\|20 | ···················· | ···················· | ····················· | ···················· | ···················· | ···················· | |
| T | 7189 | hCoV-19/England/QEUH-B66BC0/2020\|EPI_ISL_662351\|20 | ···················· | ···················· | ····················· | ···················· | ···················· | ···················· | |
| T | 7190 | hCoV-19/England/QEUH-B6A4FE/2020\|EPI_ISL_662353\|20 | ···················· | ···················· | ····················· | ···················· | ···················· | ···················· | |
| T | 7191 | hCoV-19/England/ALDP-B7350E/2020\|EPI_ISL_662354\|20 | ···················· | ···················· | ····················· | ···················· | ···················· | ···················· | |
| T | 7192 | hCoV-19/England/QEUH-B6AA1A/2020\|EPI_ISL_662355\|20 | ···················· | ···················· | ····················· | ···················· | ···················· | ···················· | |
| T | 7193 | hCoV-19/England/QEUH-B6ADB7/2020\|EPI_ISL_662357\|20 | ···················· | ···················· | ····················· | ···················· | ···················· | ···················· | |
| T | 7194 | hCoV-19/England/ALDP-B7393F/2020\|EPI_ISL_662359\|20 | ···················· | ···················· | ····················· | ···················· | ···················· | ···················· | |
| T | 7195 | hCoV-19/England/QEUH-B6A5FB/2020\|EPI_ISL_662361\|20 | ···················· | ···················· | ····················· | ···················· | ···················· | ···················· | |
| T | 7196 | hCoV-19/England/QEUH-B6A607/2020\|EPI_ISL_662362\|20 | ···················· | ···················· | ····················· | ···················· | ···················· | ···················· | |
| T | 7197 | hCoV-19/England/QEUH-B66911/2020\|EPI_ISL_662364\|20 | ···················· | ···················· | ····················· | ···················· | ···················· | ···················· | |
| T | 7198 | hCoV-19/England/ALDP-B736ED/2020\|EPI_ISL_662365\|20 | ···················· | ···················· | ····················· | ···················· | ···················· | ···················· | |
| T | 7199 | hCoV-19/England/QEUH-B6ADE4/2020\|EPI_ISL_662366\|20 | ···················· | ···················· | ····················· | ···················· | ···················· | ···················· | |
| T | 7200 | hCoV-19/England/QEUH-B675C1/2020\|EPI_ISL_662367\|20 | ···················· | ···················· | ····················· | ···················· | ···················· | ···················· | |
| T | 7201 | hCoV-19/England/QEUH-B6A7B9/2020\|EPI_ISL_662368\|20 | ···················· | ···················· | ····················· | ···················· | ···················· | ···················· | |
| T | 7202 | hCoV-19/England/ALDP-B7388D/2020\|EPI_ISL_662369\|20 | ···················· | ···················· | ····················· | ···················· | ···················· | ···················· | |
| T | 7203 | hCoV-19/England/QEUH-B66832/2020\|EPI_ISL_662370\|20 | ···················· | ···················· | ····················· | ···················· | ···················· | ···················· | |
| T | 7204 | hCoV-19/England/QEUH-B6727F/2020\|EPI_ISL_662371\|20 | ···················· | ···················· | ····················· | ···················· | ······T············· | ···················· | |
| T | 7205 | hCoV-19/England/ALDP-B73780/2020\|EPI_ISL_662374\|20 | ··················T· | ···················· | ····················· | ···················· | ···················· | ···················· | 1 |
| T | 7206 | hCoV-19/England/ALDP-B736B0/2020\|EPI_ISL_662375\|20 | ···················· | ···················· | ····················· | ···················· | ···················· | ···················· | |
| T | 7207 | hCoV-19/England/QEUH-B6A731/2020\|EPI_ISL_662376\|20 | ···················· | ···················· | ····················· | ···················· | ···················· | ···················· | |

T 7208 hCoV-19/England/QEUH-B6ADD5/2020|EPI_ISL_662377|20 ···················· ···················· ···················· ···················· ···················· ····················
T 7209 hCoV-19/Australia/VIC16841/2020|EPI_ISL_663600|202 ···················· ···················· ···················· ···················· ···················· ····················
T 7210 hCoV-19/Australia/VIC16843/2020|EPI_ISL_663601|202 ···················· ···················· ···················· ···················· ···················· ····················
T 7211 hCoV-19/Australia/VIC16846/2020|EPI_ISL_663602|202 ···················· ···················· ···················· ···················· ···················· ····················
T 7212 hCoV-19/Australia/VIC16848/2020|EPI_ISL_663603|202 ···················· ···················· ···················· ···················· ···················· ····················
T 7213 hCoV-19/Australia/VIC16850/2020|EPI_ISL_663604|202 ···················· ···················· ···················· ···················· ···················· ····················
T 7214 hCoV-19/Australia/VIC16852/2020|EPI_ISL_663605|202 ···················· ···················· ···················· ···················· ···················· ····················
T 7215 hCoV-19/Australia/VIC16853/2020|EPI_ISL_663606|202 ···················· ···················· ···················· ···················· ···················· ····················
T 7216 hCoV-19/Australia/VIC16854/2020|EPI_ISL_663607|202 ···················· ···················· ···················· ···················· ···················· ····················
T 7217 hCoV-19/Australia/VIC16855/2020|EPI_ISL_663608|202 ···················· ···················· ···················· ···················· ···················· ····················
T 7218 hCoV-19/Australia/VIC16857/2020|EPI_ISL_663609|202 ···················· ···················· ···················· ···················· ···················· ····················
T 7219 hCoV-19/Australia/VIC16858/2020|EPI_ISL_663610|202 ···················· ···················· ···················· ···················· ···················· NN··················
T 7221 hCoV-19/Australia/VIC16863/2020|EPI_ISL_663612|202 ···················· ···················· ···················· ···················· ···················· ····················
T 7223 hCoV-19/Australia/VIC16869/2020|EPI_ISL_663614|202 ···················· ···················· ···················· ···················· ···················· ····················
T 7225 hCoV-19/Australia/VIC16874/2020|EPI_ISL_663616|202 ···················· ···················· ···················· ···················· ···················· ····················
T 7226 hCoV-19/Australia/VIC16875/2020|EPI_ISL_663617|202 ···················· ···················· ···················· ···················· ···················· ····················
T 7227 hCoV-19/Australia/VIC16879/2020|EPI_ISL_663618|202 ···················· ···················· ···················· ···················· ···················· ····················
T 7230 hCoV-19/Australia/VIC16944/2020|EPI_ISL_663654|202 ···················· ···················· ···················· ···················· ···················· ····················
T 7231 hCoV-19/Australia/VIC16945/2020|EPI_ISL_663655|202 ···················· ···················· ···················· ···················· ···················· ····················
T 7232 hCoV-19/Australia/VIC16946/2020|EPI_ISL_663656|202 ···················· ···················· ···················· ···················· ···················· ····················
T 7234 hCoV-19/Australia/VIC16968/2020|EPI_ISL_663665|202 ···················· ···················· ···················· ···················· ···················· ····················
T 7235 hCoV-19/Australia/VIC16970/2020|EPI_ISL_663666|202 ···················· ···················· ···················· ···················· ···················· ····················
T 7238 hCoV-19/Australia/VIC16979/2020|EPI_ISL_663669|202 ···················· ···················· ···················· ···················· ···················· ····················
T 7239 hCoV-19/Australia/VIC16983/2020|EPI_ISL_663670|202 ···················· ···················· ···················· ···················· ···················· ····················
T 7241 hCoV-19/Australia/VIC16986/2020|EPI_ISL_663672|202 ···················· ···················· ···················· ···················· ···················· ····················
T 7243 hCoV-19/Australia/VIC16989/2020|EPI_ISL_663674|202 ···················· ···················· ···················· ···················· ···················· ····················
T 7244 hCoV-19/Australia/VIC16994/2020|EPI_ISL_663675|202 ···················· ···················· ···················· ···················· ···················· ····················
T 7245 hCoV-19/Australia/VIC16996/2020|EPI_ISL_663676|202 ···················· ···················· ···················· ···················· ···················· ····················
T 7246 hCoV-19/Australia/VIC16997/2020|EPI_ISL_663677|202 ···················· ···················· ···················· ···················· ···················· ····················
T 7247 hCoV-19/Australia/VIC16998/2020|EPI_ISL_663678|202 ···················· ···················· ···················· ···················· ···················· ····················
T 7248 hCoV-19/Australia/VIC16999/2020|EPI_ISL_663679|202 ···················· ···················· ···················· ···················· ···················· ····················
T 7249 hCoV-19/Australia/VIC17001/2020|EPI_ISL_663680|202 ···················· ···················· ···················· ···················· ···················· ····················
T 7250 hCoV-19/Australia/VIC17004/2020|EPI_ISL_663681|202 ···················· ···················· ···················· ···················· ···················· ····················
T 7251 hCoV-19/Australia/VIC17005/2020|EPI_ISL_663682|202 ···················· ···················· ···················· ···················· ···················· ····················
T 7252 hCoV-19/Australia/VIC17006/2020|EPI_ISL_663683|202 ···················· ···················· ···················· ···················· ···················· ····················
T 7253 hCoV-19/Australia/VIC17007/2020|EPI_ISL_663684|202 ···················· ···················· ···················· ···················· ···················· ····················
T 7254 hCoV-19/Australia/VIC17010/2020|EPI_ISL_663685|202 ···················· ···················· ···················· ···················· ···················· ····················
T 7255 hCoV-19/Australia/VIC17019/2020|EPI_ISL_663686|202 ···················· ···················· ···················· ···················· ···················· ····················
T 7256 hCoV-19/Australia/VIC17020/2020|EPI_ISL_663687|202 ···················· ···················· ···················· ···················· ···················· ····················
T 7257 hCoV-19/Australia/VIC17021/2020|EPI_ISL_663688|202 ···················· ···················· ···················· ···················· ···················· ····················
T 7259 hCoV-19/Australia/VIC17025/2020|EPI_ISL_663690|202 ···················· ···················· ···················· ···················· ···················· ····················
T 7260 hCoV-19/Australia/VIC17026/2020|EPI_ISL_663691|202 ···················· ···················· ···················· ···················· ···················· ····················
T 7261 hCoV-19/Australia/VIC17028/2020|EPI_ISL_663692|202 ···················· ···················· ···················· ···················· ···················· ····················
T 7262 hCoV-19/Australia/VIC17029/2020|EPI_ISL_663693|202 ···················· ···················· ···················· ···················· ···················· ····················
T 7263 hCoV-19/Australia/VIC17031/2020|EPI_ISL_663694|202 ···················· ···················· ···················· ···················· ···················· ····················
T 7264 hCoV-19/Australia/VIC17033/2020|EPI_ISL_663695|202 ···················· ···················· ···················· ···················· ···················· ····················
T 7265 hCoV-19/Australia/VIC17037/2020|EPI_ISL_663696|202 ···················· ···················· ···················· ···················· ···················· ····················
T 7266 hCoV-19/Australia/VIC17040/2020|EPI_ISL_663697|202 ···················· ···················· ···················· ···················· ···················· ····················
T 7267 hCoV-19/Australia/VIC17041/2020|EPI_ISL_663698|202 ···················· ···················· ···················· ···················· ···················· ····················
T 7268 hCoV-19/Australia/VIC17042/2020|EPI_ISL_663699|202 ···················· ···················· ···················· ···················· ···················· ····················
T 7269 hCoV-19/England/ALDP-B74118/2020|EPI_ISL_664900|20 ···················· ···················· ···················· ···················· ···················· N···················
T 7270 hCoV-19/England/ALDP-B74136/2020|EPI_ISL_664901|20 ···················· ···················· ···················· ···················· ···················· ····················
T 7272 hCoV-19/England/ALDP-B74163/2020|EPI_ISL_664903|20 ···················· ···················· ···················· ···················· ···················· ····················
T 7273 hCoV-19/England/ALDP-B74172/2020|EPI_ISL_664904|20 ···················· ···················· ···················· ···················· ···················· ····················
T 7274 hCoV-19/England/ALDP-B74181/2020|EPI_ISL_664905|20 ···················· ···················· ···················· ···················· ···················· ····················
T 7275 hCoV-19/England/ALDP-B74190/2020|EPI_ISL_664906|20 ···················· ···················· ···················· ···················· ···················· ····················
T 7277 hCoV-19/England/ALDP-B741BE/2020|EPI_ISL_664908|20 ···················· ···················· ···················· ···················· ···················· ····················
T 7278 hCoV-19/England/ALDP-B741CD/2020|EPI_ISL_664909|20 ···················· ········T··········· ···················· ···················· ···················· ····················
T 7280 hCoV-19/England/ALDP-B74215/2020|EPI_ISL_664911|20 ···················· ···················· ···················· ···················· ···················· ····················
T 7281 hCoV-19/England/ALDP-B7427F/2020|EPI_ISL_664912|20 ···················· ···················· ···················· ···················· NN·················· ····················
T 7282 hCoV-19/England/ALDP-B743E5/2020|EPI_ISL_664913|20 ···················· ···················· ···················· ···················· ···················· ····················
T 7283 hCoV-19/England/ALDP-B74F1E/2020|EPI_ISL_664914|20 ···················· ···················· ···················· ···················· ···················· ····················
T 7284 hCoV-19/England/ALDP-B74F2D/2020|EPI_ISL_664915|20 ···················· ···················· ···················· ···················· ···················· ····················
T 7285 hCoV-19/England/ALDP-B74F3C/2020|EPI_ISL_664916|20 ···················· ···················· ···················· ···················· ···················· ····················
T 7286 hCoV-19/England/ALDP-B74F4B/2020|EPI_ISL_664917|20 ···················· ···················· ···················· ···················· ···················· ····················
T 7287 hCoV-19/England/ALDP-B74F5A/2020|EPI_ISL_664918|20 ···················· ···················· ···················· ···················· ·····T·············· ····················
T 7289 hCoV-19/England/ALDP-B74FA5/2020|EPI_ISL_664920|20 ···················· ···················· ···················· ···················· ···················· ····················
T 7290 hCoV-19/England/ALDP-B74FC3/2020|EPI_ISL_664921|20 ···················· ···················· ···················· ···················· ···················· ····················
T 7291 hCoV-19/England/ALDP-B753B7/2020|EPI_ISL_664922|20 ···················· ···················· ···················· ···················· ···················· ····················
T 7292 hCoV-19/England/ALDP-B753D5/2020|EPI_ISL_664923|20 ···················· ···················· ···················· ···················· ···················· ····················
T 7293 hCoV-19/England/ALDP-B753E4/2020|EPI_ISL_664924|20 ···················· ···················· ···················· ···················· ···················· ····················
T 7294 hCoV-19/England/ALDP-B753F3/2020|EPI_ISL_664925|20 ···················· ···················· ···················· ···················· ···················· ····················
T 7295 hCoV-19/England/ALDP-B7541E/2020|EPI_ISL_664926|20 ···················· ···················· ···················· ···················· ···················· ····················
T 7296 hCoV-19/England/ALDP-B75A1C/2020|EPI_ISL_664927|20 ···················· ···················· ···················· ···················· ···················· ····················
T 7297 hCoV-19/England/ALDP-B75A3A/2020|EPI_ISL_664928|20 ···················· ···················· ···················· ···················· ···················· ····················
T 7298 hCoV-19/England/ALDP-B75A49/2020|EPI_ISL_664929|20 ···················· ···················· ···················· ···················· N··················· ····················
T 7300 hCoV-19/England/ALDP-B75A85/2020|EPI_ISL_664931|20 ···················· ···················· ···················· ···················· ···················· ····················
T 7301 hCoV-19/England/ALDP-B75A94/2020|EPI_ISL_664932|20 ···················· ···················· ···················· ···················· ···················· ····················
T 7303 hCoV-19/England/ALDP-B75AEF/2020|EPI_ISL_664934|20 ···················· ···················· ···················· ···················· ···················· ····················
T 7304 hCoV-19/England/ALDP-B772F4/2020|EPI_ISL_664935|20 ···················· ···················· ···················· ···················· ···················· ····················

| | | | | | | | |
|---|---|---|---|---|---|---|---|
| T | 7305 hCoV-19/England/ALDP-B77300/2020\|EPI_ISL_664936\|20 | ···················· | ·A·················· | ····················· | ···················· | ···················· | ···A················ |
| T | 7306 hCoV-19/England/ALDP-B7731F/2020\|EPI_ISL_664937\|20 | ···················· | ···················· | ····················· | ···················· | ···················· | ···················· |
| T | 7307 hCoV-19/England/ALDP-B7732E/2020\|EPI_ISL_664938\|20 | ···················· | ···················· | ····················· | ···················· | ···················· | ···················· |
| T | 7310 hCoV-19/England/CAMB-1B95E8/2020\|EPI_ISL_664941\|20 | ···················· | ···················· | ····················· | ···················· | ···················· | ···················· |
| T | 7311 hCoV-19/England/CAMB-1B95F7/2020\|EPI_ISL_664942\|20 | ···················· | ···················· | ····················· | ···················· | ···················· | ···················· |
| T | 7312 hCoV-19/England/CAMB-1B973D/2020\|EPI_ISL_664943\|20 | ···················· | ···················· | ····················· | ···················· | ···················· | ···················· |
| T | 7313 hCoV-19/England/CAMB-1B9788/2020\|EPI_ISL_664944\|20 | ···················· | ···················· | ····················· | ···················· | ···················· | ···················· |
| T | 7314 hCoV-19/England/CAMB-1B97E2/2020\|EPI_ISL_664945\|20 | ···················· | ···················· | ····················· | ···················· | ···················· | ···················· |
| T | 7315 hCoV-19/England/CAMB-1B98B2/2020\|EPI_ISL_664946\|20 | ···················· | ···················· | ····················· | ···················· | ···················· | ···················· |
| T | 7316 hCoV-19/England/CAMB-1B98C1/2020\|EPI_ISL_664947\|20 | ···················· | ···················· | ····················· | ···················· | ···················· | ···················· |
| T | 7317 hCoV-19/England/CAMB-1B98FE/2020\|EPI_ISL_664948\|20 | ···················· | ···················· | ····················· | ···················· | ···················· | ···················· |
| T | 7318 hCoV-19/England/CAMB-1B990A/2020\|EPI_ISL_664949\|20 | ···················· | ···················· | ····················· | ···················· | ···················· | ···················· |
| T | 7319 hCoV-19/England/CAMB-1B9919/2020\|EPI_ISL_664950\|20 | ···················· | ···················· | ····················· | ···················· | ···················· | ···················· |
| T | 7320 hCoV-19/England/CAMB-1B9928/2020\|EPI_ISL_664951\|20 | ···················· | ···················· | ····················· | ···················· | ···················· | ···················· |
| T | 7321 hCoV-19/England/CAMB-1BA4CC/2020\|EPI_ISL_664985\|20 | ···················· | ···················· | ····················· | ···················· | ···················· | ···A················ |
| T | 7322 hCoV-19/England/CAMB-1BA4DB/2020\|EPI_ISL_664986\|20 | ···················· | ···················· | ····················· | ···················· | ···················· | ···················· |
| T | 7323 hCoV-19/England/CAMB-1BA4EA/2020\|EPI_ISL_664987\|20 | ···················· | ···················· | ····················· | ···················· | ···················· | ···················· |
| T | 7324 hCoV-19/England/CAMB-1BA58D/2020\|EPI_ISL_664988\|20 | ···················· | ···················· | ····················· | ···················· | ···················· | ···················· |
| T | 7325 hCoV-19/England/CAMB-1BA7A5/2020\|EPI_ISL_664996\|20 | ···················· | ···················· | ····················· | ···················· | ···················· | ···················· |
| T | 7326 hCoV-19/England/CAMB-1BA7C3/2020\|EPI_ISL_664997\|20 | ···················· | ···················· | ····················· | ···················· | ···················· | ···················· |
| T | 7327 hCoV-19/England/CAMB-1BAD2B/2020\|EPI_ISL_664998\|20 | ···················· | ···················· | ····················· | ···················· | ···················· | ···················· |
| T | 7328 hCoV-19/England/CAMB-1BAE82/2020\|EPI_ISL_664999\|20 | ···················· | ·A·················· | ····················· | ···················· | ···················· | ···A················ |
| T | 7329 hCoV-19/USA/OR-OHSU-2219/2020\|EPI_ISL_667150\|2020- | ···················· | ···················· | ····················· | ···················· | ···················· | ···················· |
| T | 7330 hCoV-19/USA/OR-OHSU-2221/2020\|EPI_ISL_667151\|2020- | ···················· | ···················· | ····················· | ···················· | ···················· | ···················· |
| T | 7331 hCoV-19/USA/OR-OHSU-2225/2020\|EPI_ISL_667152\|2020- | ···················· | ···················· | ····················· | ···················· | ···················· | ···················· |
| T | 7332 hCoV-19/USA/OR-OHSU-2227/2020\|EPI_ISL_667153\|2020- | ···················· | ···················· | ····················· | ···················· | ···················· | ···················· |
| T | 7333 hCoV-19/USA/OR-OHSU-2233/2020\|EPI_ISL_667154\|2020- | ···················· | ···A················ | ····················· | ···················· | ···················· | ···················· |
| T | 7334 hCoV-19/USA/OR-OHSU-2239/2020\|EPI_ISL_667156\|2020- | ···················· | ···················· | ····················· | ···················· | ···················· | ···················· |
| T | 7335 hCoV-19/USA/OR-OHSU-2241/2020\|EPI_ISL_667157\|2020- | ···················· | ···················· | ····················· | ···················· | ···················· | ···················· |
| T | 7336 hCoV-19/USA/OR-OHSU-2249/2020\|EPI_ISL_667158\|2020- | ···················· | ···················· | ····················· | ···················· | ···················· | ···················· |
| T | 7337 hCoV-19/USA/OR-OHSU-2251/2020\|EPI_ISL_667159\|2020- | ···················· | ···················· | ····················· | ···················· | ···················· | ···················· |
| T | 7338 hCoV-19/USA/OR-OHSU-2253/2020\|EPI_ISL_667160\|2020- | ···················· | ···················· | ····················· | ···················· | ···················· | ···················· |
| T | 7339 hCoV-19/USA/OR-OHSU-2255/2020\|EPI_ISL_667161\|2020- | ···················· | ·A·················· | ····················· | ···················· | ···················· | ···················· |
| T | 7340 hCoV-19/USA/OR-OHSU-2257/2020\|EPI_ISL_667162\|2020- | ···················· | ···················· | ····················· | ···················· | ···················· | ···················· |
| T | 7341 hCoV-19/USA/OR-OHSU-2261/2020\|EPI_ISL_667163\|2020- | ···················· | ···················· | ····················· | ···················· | ···················· | ···················· |
| T | 7342 hCoV-19/USA/OR-OHSU-2269/2020\|EPI_ISL_667164\|2020- | ···················· | ···················· | ····················· | ···················· | ···················· | ···················· |
| T | 7343 hCoV-19/USA/FL-OHSU-2271/2020\|EPI_ISL_667165\|2020- | ···················· | ···················· | ····················· | ···················· | ···················· | ···················· |
| T | 7344 hCoV-19/USA/OR-OHSU-2273/2020\|EPI_ISL_667166\|2020- | ···················· | ···················· | ····················· | ···················· | ···················· | ···················· |
| T | 7345 hCoV-19/USA/OR-OHSU-2275/2020\|EPI_ISL_667167\|2020- | ···················· | ···················· | ····················· | ···················· | ···················· | ···················· |
| T | 7346 hCoV-19/USA/OR-OHSU-2277/2020\|EPI_ISL_667168\|2020- | ···················· | ···················· | ····················· | ···················· | ···················· | ···················· |
| T | 7347 hCoV-19/USA/WA-OHSU-2287/2020\|EPI_ISL_667169\|2020- | ···················· | ·A·················· | ····················· | ···················· | ···················· | ···················· |
| T | 7348 hCoV-19/USA/OR-OHSU-2289/2020\|EPI_ISL_667170\|2020- | ···················· | ···················· | ····················· | ···················· | ···················· | ···················· |
| T | 7349 hCoV-19/USA/OR-OHSU-2291/2020\|EPI_ISL_667171\|2020- | ···················· | ···················· | ····················· | ···················· | ···················· | ···················· |
| T | 7350 hCoV-19/USA/OR-OHSU-2295/2020\|EPI_ISL_667172\|2020- | ···················· | ·A·················· | ····················· | ···················· | ···················· | ···················· |
| T | 7351 hCoV-19/USA/OR-OHSU-2297/2020\|EPI_ISL_667173\|2020- | ···················· | ···················· | ····················· | ···················· | ···················· | ···················· |
| T | 7352 hCoV-19/USA/OR-OHSU-2299/2020\|EPI_ISL_667174\|2020- | ···················· | ·A·················· | ····················· | ···················· | ···················· | ···················· |
| T | 7353 hCoV-19/USA/OR-OHSU-2301/2020\|EPI_ISL_667175\|2020- | ···················· | ·A·················· | ····················· | ···················· | ···················· | ···················· |
| T | 7354 hCoV-19/USA/OR-OHSU-2303/2020\|EPI_ISL_667176\|2020- | ···················· | ·A·················· | ····················· | ···················· | ···················· | ···················· |
| T | 7355 hCoV-19/USA/OR-OHSU-2305/2020\|EPI_ISL_667177\|2020- | ···················· | ···················· | ····················· | ···················· | ···················· | ···················· |
| T | 7356 hCoV-19/USA/OR-OHSU-2307/2020\|EPI_ISL_667178\|2020- | ···················· | ···················· | ····················· | ···················· | ···················· | ···················· |
| T | 7357 hCoV-19/USA/OR-OHSU-2309/2020\|EPI_ISL_667179\|2020- | ···················· | ·············A······ | ····················· | ···················· | ···················· | ···················· |
| T | 7358 hCoV-19/USA/OR-OHSU-2313/2020\|EPI_ISL_667180\|2020- | ···················· | ···················· | ····················· | ···················· | ···················· | ···················· |
| T | 7359 hCoV-19/USA/OR-OHSU-2319/2020\|EPI_ISL_667181\|2020- | ···················· | ···················· | ····················· | ···················· | ···················· | ···················· |
| T | 7360 hCoV-19/USA/OR-OHSU-2321/2020\|EPI_ISL_667182\|2020- | ···················· | ···················· | ····················· | ···················· | ···················· | ···················· |
| T | 7361 hCoV-19/USA/OR-OHSU-2329/2020\|EPI_ISL_667184\|2020- | ···················· | ···················· | ····················· | ···················· | ···················· | ···················· |
| T | 7362 hCoV-19/USA/OR-OHSU-2331/2020\|EPI_ISL_667185\|2020- | ···················· | ···················· | ····················· | ···················· | ···················· | ···················· |
| T | 7363 hCoV-19/USA/WA-OHSU-2337/2020\|EPI_ISL_667187\|2020- | ···················· | ···················· | ····················· | ···················· | ···················· | ···················· |
| T | 7364 hCoV-19/USA/OR-OHSU-2339/2020\|EPI_ISL_667188\|2020- | ···················· | ·A·················· | ····················· | ···················· | ···················· | ···················· |
| T | 7365 hCoV-19/USA/OR-OHSU-2341/2020\|EPI_ISL_667189\|2020- | ···················· | ···················· | ····················· | ···················· | ···················· | ···················· |
| T | 7366 hCoV-19/USA/OR-OHSU-2345/2020\|EPI_ISL_667190\|2020- | ···················· | ···················· | ····················· | ···················· | ···················· | ···················· |
| T | 7367 hCoV-19/USA/OR-OHSU-2353/2020\|EPI_ISL_667191\|2020- | ···················· | ···················· | ····················· | ···················· | ···················· | ···················· |
| T | 7368 hCoV-19/USA/OR-OHSU-2361/2020\|EPI_ISL_667192\|2020- | ···················· | ···················· | ····················· | ···················· | ···················· | ···················· |
| T | 7369 hCoV-19/USA/OR-OHSU-2365/2020\|EPI_ISL_667193\|2020- | ···················· | ·A·················· | ····················· | ···················· | ···················· | ···················· |
| T | 7370 hCoV-19/USA/OR-OHSU-2369/2020\|EPI_ISL_667194\|2020- | ···················· | ···················· | ····················· | ···················· | ···················· | ···················· |
| T | 7371 hCoV-19/USA/OR-OHSU-2373/2020\|EPI_ISL_667195\|2020- | ···················· | ···················· | ····················· | ···················· | ···················· | ···················· |
| T | 7372 hCoV-19/USA/OR-OHSU-2395/2020\|EPI_ISL_667196\|2020- | ···················· | ···················· | ····················· | ···················· | ···················· | ···················· |
| T | 7373 hCoV-19/USA/OR-OHSU-2397/2020\|EPI_ISL_667197\|2020- | ···················· | ···················· | ····················· | ···················· | ···················· | ···················· |
| T | 7374 hCoV-19/USA/OR-OHSU-2409/2020\|EPI_ISL_667199\|2020- | ···················· | ···················· | ····················· | ···················· | ···················· | ···················· |
| T | 7375 hCoV-19/USA/OR-OHSU-2411/2020\|EPI_ISL_667200\|2020- | ···················· | ···················· | ····················· | ···················· | ···················· | ···················· |
| T | 7376 hCoV-19/USA/OR-OHSU-2419/2020\|EPI_ISL_667201\|2020- | ···················· | ···A················ | ····················· | ···················· | ···················· | ···················· |
| T | 7377 hCoV-19/USA/OR-OHSU-2421/2020\|EPI_ISL_667202\|2020- | ···················· | ···················· | ····················· | ···················· | ···················· | ···················· |
| T | 7378 hCoV-19/USA/OR-OHSU-2423/2020\|EPI_ISL_667203\|2020- | ···················· | ·A·················· | ····················· | ···················· | ···················· | ···················· |
| T | 7379 hCoV-19/USA/OR-OHSU-2429/2020\|EPI_ISL_667205\|2020- | ···················· | ·A·················· | ····················· | ···················· | ···················· | ···················· |
| T | 7380 hCoV-19/Denmark/DCGC-10232/2020\|EPI_ISL_669847\|202 | ···················· | ···················· | ····················· | ···················· | ···················· | ···················· |
| T | 7381 hCoV-19/Denmark/DCGC-10233/2020\|EPI_ISL_669848\|202 | ···················· | ···················· | ····················· | ···················· | ···················· | ···················· |
| T | 7382 hCoV-19/Denmark/DCGC-10234/2020\|EPI_ISL_669849\|202 | ···················· | ···················· | ····················· | ···················· | ···················· | ···················· |
| T | 7383 hCoV-19/Denmark/DCGC-10235/2020\|EPI_ISL_669850\|202 | ···················· | ···················· | ····················· | ···················· | ···················· | ···················· |
| T | 7384 hCoV-19/Denmark/DCGC-10238/2020\|EPI_ISL_669851\|202 | ···················· | ···················· | ····················· | ···················· | ···················· | ···················· |
| T | 7385 hCoV-19/Denmark/DCGC-10239/2020\|EPI_ISL_669852\|202 | ···················· | ·A·················· | ····················· | ···················· | ···················· | ···A················ |
| T | 7386 hCoV-19/Denmark/DCGC-10241/2020\|EPI_ISL_669853\|202 | ···················· | ···················· | ····················· | ···················· | ···················· | ···················· |

| | | | | | | | |
|---|---|---|---|---|---|---|---|
| T | 7387 hCoV-19/Denmark/DCGC-10244/2020\|EPI_ISL_669854\|202 | ···················· | ···················· | ····················· | ···················· | ···················· | ···················· |
| T | 7388 hCoV-19/Denmark/DCGC-10245/2020\|EPI_ISL_669855\|202 | ···················· | ···················· | ····················· | ···················· | ···················· | ···················· |
| T | 7389 hCoV-19/Denmark/DCGC-10246/2020\|EPI_ISL_669856\|202 | ···················· | ···················· | ····················· | ···················· | ···················· | ···················· |
| T | 7390 hCoV-19/Denmark/DCGC-10250/2020\|EPI_ISL_669857\|202 | ···················· | ···················· | ····················· | ···················· | ···················· | ···················· |
| T | 7391 hCoV-19/Denmark/DCGC-10254/2020\|EPI_ISL_669860\|202 | ···················· | ···················· | ····················· | ···················· | ···················· | ···················· |
| T | 7392 hCoV-19/Denmark/DCGC-10256/2020\|EPI_ISL_669861\|202 | ···················· | ···················· | ····················· | ···················· | ···················· | ···················· |
| T | 7393 hCoV-19/Denmark/DCGC-10257/2020\|EPI_ISL_669862\|202 | ···················· | ···················· | ····················· | ···················· | ·····T·············· | ···················· |
| T | 7394 hCoV-19/Denmark/DCGC-10258/2020\|EPI_ISL_669863\|202 | ···················· | ···················· | ····················· | ···················· | ···················· | ···················· |
| T | 7395 hCoV-19/Denmark/DCGC-10260/2020\|EPI_ISL_669864\|202 | ···················· | ···················· | ····················· | ···················· | ···················· | ···················· |
| T | 7396 hCoV-19/Denmark/DCGC-10262/2020\|EPI_ISL_669865\|202 | ···················· | ···················· | ····················· | ···················· | ···················· | ···················· |
| T | 7397 hCoV-19/Denmark/DCGC-10265/2020\|EPI_ISL_669866\|202 | ···················· | ···················· | ····················· | ···················· | ···················· | ···················· |
| T | 7398 hCoV-19/Denmark/DCGC-10266/2020\|EPI_ISL_669867\|202 | ···················· | ···················· | ····················· | ···················· | ···················· | ···················· |
| T | 7399 hCoV-19/Denmark/DCGC-10271/2020\|EPI_ISL_669868\|202 | ···················· | ···················· | ····················· | ···················· | ···················· | ···················· |
| T | 7400 hCoV-19/Denmark/DCGC-11248/2020\|EPI_ISL_670793\|202 | ···················· | ···················· | ····················· | ···················· | ···················· | ···················· |
| T | 7401 hCoV-19/Denmark/DCGC-11252/2020\|EPI_ISL_670794\|202 | ···················· | ···················· | ····················· | ···················· | ···················· | ···················· |
| T | 7402 hCoV-19/Denmark/DCGC-11254/2020\|EPI_ISL_670795\|202 | ···················· | ···················· | ····················· | ···················· | ···················· | ···················· |
| T | 7403 hCoV-19/Denmark/DCGC-11256/2020\|EPI_ISL_670796\|202 | ···················· | ···················· | ····················· | ···················· | ···················· | ···················· |
| T | 7404 hCoV-19/Denmark/DCGC-11257/2020\|EPI_ISL_670797\|202 | ···················· | ···················· | ····················· | ···················· | ···················· | ···················· |
| T | 7405 hCoV-19/Denmark/DCGC-11258/2020\|EPI_ISL_670798\|202 | ···················· | ···················· | ····················· | ···················· | ···················· | ···················· |
| T | 7406 hCoV-19/Denmark/DCGC-11260/2020\|EPI_ISL_670799\|202 | ···················· | ···················· | ····················· | ···················· | ···················· | ···················· |
| T | 7407 hCoV-19/Denmark/DCGC-11308/2020\|EPI_ISL_670830\|202 | ···················· | ···················· | ····················· | ···················· | ···················· | ···················· |
| T | 7408 hCoV-19/Denmark/DCGC-11309/2020\|EPI_ISL_670831\|202 | ···················· | ···················· | ····················· | ···················· | ···················· | ···················· |
| T | 7409 hCoV-19/Denmark/DCGC-11314/2020\|EPI_ISL_670832\|202 | ···················· | ···················· | ····················· | ···················· | ···················· | ···················· |
| T | 7410 hCoV-19/Denmark/DCGC-11316/2020\|EPI_ISL_670833\|202 | ···················· | ···················· | ····················· | ···················· | ···················· | ···················· |
| T | 7411 hCoV-19/Denmark/DCGC-11317/2020\|EPI_ISL_670834\|202 | ···················· | ···················· | ····················· | ···················· | ···················· | ···················· |
| T | 7412 hCoV-19/Denmark/DCGC-11318/2020\|EPI_ISL_670835\|202 | ···················· | ···················· | ····················· | ···················· | ···················· | ···················· |
| T | 7413 hCoV-19/Denmark/DCGC-11319/2020\|EPI_ISL_670836\|202 | ···················· | ···················· | ····················· | ···················· | ···················· | ···················· |
| T | 7414 hCoV-19/Denmark/DCGC-11320/2020\|EPI_ISL_670837\|202 | ···················· | ···················· | ····················· | ···················· | ···················· | ···················· |
| T | 7415 hCoV-19/Denmark/DCGC-11321/2020\|EPI_ISL_670838\|202 | ···················· | ·A·················· | ····················· | ···················· | ···················· | ···················· |
| T | 7416 hCoV-19/Denmark/DCGC-11322/2020\|EPI_ISL_670839\|202 | ···················· | ···················· | ····················· | ···················· | ···················· | ···················· |
| T | 7417 hCoV-19/Denmark/DCGC-11324/2020\|EPI_ISL_670840\|202 | ···················· | ···················· | ····················· | ···················· | ···················· | ···················· |
| T | 7418 hCoV-19/Denmark/DCGC-11325/2020\|EPI_ISL_670841\|202 | ···················· | ···················· | ····················· | ···················· | ···················· | ···················· |
| T | 7419 hCoV-19/Denmark/DCGC-11326/2020\|EPI_ISL_670842\|202 | ···················· | ···················· | ····················· | ···················· | ···················· | ···················· |
| T | 7420 hCoV-19/Denmark/DCGC-11327/2020\|EPI_ISL_670843\|202 | ···················· | ·A·················· | ····················· | ···················· | ···················· | ···················· |
| T | 7421 hCoV-19/Denmark/DCGC-11331/2020\|EPI_ISL_670844\|202 | ···················· | ···················· | ····················· | ···················· | ···················· | ···················· |
| T | 7422 hCoV-19/Denmark/DCGC-11332/2020\|EPI_ISL_670845\|202 | ···················· | ···················· | ····················· | ···················· | ···················· | ···················· |
| T | 7423 hCoV-19/Denmark/DCGC-11338/2020\|EPI_ISL_670847\|202 | ···················· | ···················· | ····················· | ···················· | ···················· | ···················· |
| T | 7424 hCoV-19/Denmark/DCGC-11339/2020\|EPI_ISL_670848\|202 | ···················· | ···················· | ····················· | ···················· | ···················· | ···················· |
| T | 7425 hCoV-19/England/QEUH-B68618/2020\|EPI_ISL_662257\|20 | ···················· | ···················· | ····················· | ···················· | ···················· | ···················· |
| T | 7426 hCoV-19/England/ALDP-B751DB/2020\|EPI_ISL_662258\|20 | ···················· | ···················· | ····················· | ···················· | ···················· | ···················· |
| T | 7427 hCoV-19/England/ALDP-B74093/2020\|EPI_ISL_662260\|20 | ···················· | ···················· | ····················· | ···················· | ···················· | ···················· |
| T | 7428 hCoV-19/England/QEUH-B67FC3/2020\|EPI_ISL_662261\|20 | ···················· | ···················· | ····················· | ···················· | ···················· | ···················· |
| T | 7429 hCoV-19/England/QEUH-B67FB4/2020\|EPI_ISL_662262\|20 | ···················· | ···················· | ····················· | ···················· | ···················· | ···················· |
| T | 7430 hCoV-19/England/ALDP-B74C9F/2020\|EPI_ISL_662263\|20 | ···················· | ···················· | ····················· | ···················· | ···················· | ···················· |
| T | 7431 hCoV-19/England/QEUH-B69125/2020\|EPI_ISL_662264\|20 | ···················· | ···················· | ····················· | ···················· | ···················· | ···················· |
| T | 7432 hCoV-19/England/ALDP-B74A1D/2020\|EPI_ISL_662265\|20 | ···················· | ···················· | ····················· | ···················· | ···················· | ···················· |
| T | 7433 hCoV-19/England/ALDP-B74D9C/2020\|EPI_ISL_662266\|20 | ···················· | ···················· | ····················· | ···················· | ···················· | ···················· |
| T | 7434 hCoV-19/England/ALDP-B749E3/2020\|EPI_ISL_662267\|20 | ···················· | ···················· | ····················· | ···················· | ···················· | ···················· |
| T | 7435 hCoV-19/England/QEUH-B68F4A/2020\|EPI_ISL_662268\|20 | ···················· | ·A·················· | ····················· | ···················· | ···················· | ··A················· |
| T | 7436 hCoV-19/England/QEUH-B68E5C/2020\|EPI_ISL_662271\|20 | ···················· | ···················· | ····················· | ···················· | ···················· | ···················· |
| T | 7437 hCoV-19/England/ALDP-B7452B/2020\|EPI_ISL_662272\|20 | ···················· | ···················· | ····················· | ···················· | ···················· | ···················· |
| T | 7438 hCoV-19/England/ALDP-B7494D/2020\|EPI_ISL_662273\|20 | ···················· | ···················· | ····················· | ···················· | ···················· | ···················· |
| T | 7439 hCoV-19/England/ALDP-B74910/2020\|EPI_ISL_662274\|20 | ···················· | ···················· | ····················· | ···················· | ···················· | ···················· |
| T | 7440 hCoV-19/England/ALDP-B74CDB/2020\|EPI_ISL_662275\|20 | ···················· | ···················· | ····················· | ···················· | ···················· | ···················· |
| T | 7441 hCoV-19/England/ALDP-B75584/2020\|EPI_ISL_662276\|20 | ···················· | ···················· | ····················· | ···················· | ···················· | ···················· |
| T | 7442 hCoV-19/England/QEUH-B66F6A/2020\|EPI_ISL_662277\|20 | ···················· | ···················· | ····················· | ···················· | ···················· | ···················· |
| T | 7443 hCoV-19/England/QEUH-B69116/2020\|EPI_ISL_662280\|20 | ···················· | ···················· | ····················· | ···················· | ···················· | ···················· |
| T | 7444 hCoV-19/England/QEUH-B691BC/2020\|EPI_ISL_662282\|20 | ···················· | ···················· | ····················· | ···················· | ···················· | ···················· |
| T | 7445 hCoV-19/England/QEUH-B691DA/2020\|EPI_ISL_662283\|20 | ···················· | ···················· | ····················· | ···················· | ···················· | ···················· |
| T | 7446 hCoV-19/England/QEUH-B68E7A/2020\|EPI_ISL_662284\|20 | ···················· | ·A·················· | ····················· | ···················· | ···················· | ···················· |
| T | 7447 hCoV-19/England/ALDP-B7435E/2020\|EPI_ISL_662285\|20 | ···················· | ···················· | ····················· | ···················· | ···················· | ···················· |
| T | 7448 hCoV-19/England/QEUH-B674B5/2020\|EPI_ISL_662287\|20 | ···················· | ···················· | ····················· | ···················· | ···················· | ···················· |
| T | 7449 hCoV-19/England/QEUH-B6819F/2020\|EPI_ISL_662288\|20 | ···················· | ···················· | ····················· | ···················· | ···················· | ···················· |
| T | 7450 hCoV-19/England/QEUH-B6A221/2020\|EPI_ISL_662289\|20 | ···················· | ···················· | ····················· | ···················· | ···················· | ···················· |
| T | 7451 hCoV-19/England/ALDP-B7351D/2020\|EPI_ISL_662290\|20 | ···················· | ···················· | ····················· | ···················· | ···················· | ···················· |
| T | 7452 hCoV-19/England/ALDP-B733D7/2020\|EPI_ISL_662292\|20 | ···················· | ···················· | ····················· | ···················· | ···················· | ···················· |
| T | 7453 hCoV-19/England/QEUH-B6A397/2020\|EPI_ISL_662293\|20 | ···················· | ·A·················· | ····················· | ···················· | ···················· | ···················· |
| T | 7454 hCoV-19/England/QEUH-B6A1AC/2020\|EPI_ISL_662294\|20 | ···················· | ···················· | ····················· | ···················· | ···················· | ···················· |
| T | 7455 hCoV-19/England/ALDP-B733E6/2020\|EPI_ISL_662296\|20 | ···················· | ···················· | ····················· | ···················· | ···················· | ···················· |
| T | 7456 hCoV-19/England/QEUH-B6B071/2020\|EPI_ISL_662297\|20 | ···················· | ···················· | ····················· | ···················· | ···················· | ···················· |
| T | 7457 hCoV-19/England/QEUH-B6A17F/2020\|EPI_ISL_662298\|20 | ···················· | ···················· | ····················· | ···················· | ···················· | ···················· |
| T | 7458 hCoV-19/England/QEUH-B6696C/2020\|EPI_ISL_662301\|20 | ···················· | ···················· | ····················· | ···················· | ···················· | ···················· |
| T | 7459 hCoV-19/England/QEUH-B6B6CA/2020\|EPI_ISL_662302\|20 | ···················· | ···················· | ····················· | ···················· | ···················· | ··A················· |
| T | 7460 hCoV-19/England/QEUH-B6737C/2020\|EPI_ISL_662304\|20 | ···················· | ···················· | ····················· | ···················· | ···················· | ···················· |
| T | 7461 hCoV-19/England/ALDP-B738F6/2020\|EPI_ISL_662305\|20 | ···················· | ···················· | ····················· | ···················· | ···················· | ···················· |
| T | 7462 hCoV-19/England/QEUH-B6693F/2020\|EPI_ISL_662306\|20 | ···················· | ···················· | ····················· | ···················· | ···················· | ···················· |
| T | 7463 hCoV-19/England/ALDP-B73735/2020\|EPI_ISL_662309\|20 | ···················· | ···················· | ····················· | ···················· | ···················· | ···················· |
| T | 7464 hCoV-19/England/QEUH-B6A0FA/2020\|EPI_ISL_662311\|20 | ···················· | ···················· | ····················· | ···················· | ···················· | ···················· |
| T | 7465 hCoV-19/England/QEUH-B6B7A9/2020\|EPI_ISL_662313\|20 | ···················· | ···················· | ····················· | ···················· | ···················· | ···················· |
| T | 7466 hCoV-19/England/QEUH-B6A4A3/2020\|EPI_ISL_662314\|20 | ···················· | ···················· | ····················· | ···················· | ···················· | ···················· |

| | | | | | | | | |
|---|---|---|---|---|---|---|---|---|
| T | 7467 hCoV-19/England/QEUH-B67312/2020\|EPI_ISL_662315\|20 | ···················· | ···················· | ···················· | ···················· | ···················· | ···················· | |
| T | 7468 hCoV-19/England/QEUH-B6ADA8/2020\|EPI_ISL_662316\|20 | ···················· | ···················· | ···················· | ···················· | ···················· | ···················· | |
| T | 7469 hCoV-19/England/QEUH-B6A6F8/2020\|EPI_ISL_662317\|20 | ···················· | ···················· | ···················· | ···················· | ···················· | ···················· | |
| T | 7470 hCoV-19/England/ALDP-B7386F/2020\|EPI_ISL_662320\|20 | ···················· | ···················· | ···················· | ···················· | ···················· | ···················· | |
| T | 7471 hCoV-19/England/QEUH-B6A388/2020\|EPI_ISL_662328\|20 | ···················· | ·A·················· | ···················· | ···················· | ···················· | ···················· | |
| T | 7472 hCoV-19/England/QEUH-B6A6BC/2020\|EPI_ISL_662329\|20 | ···················· | ···················· | ···················· | ···················· | ···················· | ···················· | |
| T | 7473 hCoV-19/England/QEUH-B6A1CA/2020\|EPI_ISL_662330\|20 | ···················· | ···················· | ···················· | ···················· | ···················· | ···················· | |
| T | 7474 hCoV-19/England/QEUH-B66C09/2020\|EPI_ISL_662331\|20 | ···················· | ···················· | ···················· | ···················· | ···················· | ···················· | |
| T | 7475 hCoV-19/England/QEUH-B67619/2020\|EPI_ISL_662332\|20 | ···················· | ···················· | ···················· | ···················· | ···················· | ···················· | |
| T | 7476 hCoV-19/England/QEUH-B6AA38/2020\|EPI_ISL_662333\|20 | ···················· | ···················· | ···················· | ···················· | ···················· | ···················· | |
| T | 7477 hCoV-19/Australia/VIC16825/2020\|EPI_ISL_663588\|202 | ···················· | ···················· | ···················· | ···················· | ···················· | ···················· | |
| T | 7478 hCoV-19/Australia/VIC16826/2020\|EPI_ISL_663589\|202 | ···················· | ···················· | ···················· | ···················· | ···················· | ···················· | |
| T | 7479 hCoV-19/Australia/VIC16827/2020\|EPI_ISL_663590\|202 | ···················· | ···················· | ···················· | ···················· | ···················· | ···················· | |
| T | 7482 hCoV-19/Australia/VIC16834/2020\|EPI_ISL_663593\|202 | ···················· | ···················· | ···················· | ···················· | ···················· | ···················· | |
| T | 7484 hCoV-19/Australia/VIC16836/2020\|EPI_ISL_663595\|202 | ···················· | ···················· | ···················· | ···················· | ···················· | ····M··············· | |
| T | 7486 hCoV-19/Australia/VIC16838/2020\|EPI_ISL_663597\|202 | ···················· | ···················· | ···················· | ···················· | NNN················· | ···················· | 1 |
| T | 7487 hCoV-19/Australia/VIC16839/2020\|EPI_ISL_663598\|202 | ···················· | ···················· | ···················· | ···················· | ···················· | ···················· | |
| T | 7488 hCoV-19/Australia/VIC16840/2020\|EPI_ISL_663599\|202 | ···················· | ···················· | ···················· | ···················· | ···················· | ···················· | |
| T | 7490 hCoV-19/Australia/VIC16883/2020\|EPI_ISL_663622\|202 | ···················· | ···················· | ···················· | ···················· | ···················· | ···················· | |
| T | 7491 hCoV-19/Australia/VIC16886/2020\|EPI_ISL_663623\|202 | ···················· | ···················· | ···················· | ···················· | ···················· | ···················· | |
| T | 7492 hCoV-19/Australia/VIC16887/2020\|EPI_ISL_663624\|202 | ···················· | ···················· | ···················· | ···················· | ···················· | ···················· | |
| T | 7493 hCoV-19/Australia/VIC16889/2020\|EPI_ISL_663625\|202 | ···················· | ···················· | ···················· | ···················· | ···················· | ···················· | |
| T | 7496 hCoV-19/Australia/VIC16894/2020\|EPI_ISL_663628\|202 | ···················· | ···················· | ···················· | ···················· | ···················· | ···················· | |
| T | 7497 hCoV-19/Australia/VIC16896/2020\|EPI_ISL_663629\|202 | ···················· | ···················· | ···················· | ···················· | ···················· | ···················· | |
| T | 7499 hCoV-19/Australia/VIC16898/2020\|EPI_ISL_663631\|202 | ···················· | ···················· | ···················· | ···················· | ···················· | ···················· | |
| T | 7500 hCoV-19/Australia/VIC16900/2020\|EPI_ISL_663632\|202 | ···················· | ···················· | ···················· | ···················· | ···················· | ···················· | |
| T | 7501 hCoV-19/Australia/VIC16901/2020\|EPI_ISL_663633\|202 | ···················· | ···················· | ···················· | ···················· | ···················· | ···················· | |
| T | 7502 hCoV-19/Australia/VIC16906/2020\|EPI_ISL_663634\|202 | ···················· | ···················· | ···················· | ···················· | ···················· | ···················· | |
| T | 7503 hCoV-19/Australia/VIC16910/2020\|EPI_ISL_663635\|202 | ···················· | ···················· | ···················· | ···················· | ···················· | ···················· | |
| T | 7505 hCoV-19/Australia/VIC16913/2020\|EPI_ISL_663637\|202 | ···················· | ···················· | ···················· | ···················· | ···················· | ···················· | |
| T | 7507 hCoV-19/Australia/VIC16915/2020\|EPI_ISL_663639\|202 | ···················· | ···················· | ···················· | ···················· | ···················· | ···················· | |
| T | 7509 hCoV-19/Australia/VIC16918/2020\|EPI_ISL_663641\|202 | ···················· | ···················· | ···················· | ···················· | ···················· | ···················· | |
| T | 7510 hCoV-19/Australia/VIC16919/2020\|EPI_ISL_663642\|202 | ···················· | ···················· | ···················· | ···················· | ···················· | ···················· | |
| T | 7511 hCoV-19/Australia/VIC16921/2020\|EPI_ISL_663643\|202 | ···················· | ···················· | ···················· | ···················· | ···················· | ···················· | |
| T | 7512 hCoV-19/Australia/VIC16929/2020\|EPI_ISL_663644\|202 | ···················· | ···················· | ···················· | ···················· | ···················· | ···················· | |
| T | 7514 hCoV-19/Australia/VIC16935/2020\|EPI_ISL_663646\|202 | ···················· | ···················· | ···················· | ···················· | ···················· | ···················· | |
| T | 7515 hCoV-19/Australia/VIC16937/2020\|EPI_ISL_663647\|202 | ···················· | ···················· | ···················· | ···················· | ···················· | ···················· | |
| T | 7516 hCoV-19/Australia/VIC16938/2020\|EPI_ISL_663648\|202 | ···················· | ···················· | ···················· | ···················· | ···················· | ···················· | |
| T | 7517 hCoV-19/Australia/VIC16939/2020\|EPI_ISL_663649\|202 | ···················· | ···················· | ···················· | ···················· | ···················· | ···················· | |
| T | 7518 hCoV-19/Australia/VIC16940/2020\|EPI_ISL_663650\|202 | ···················· | ···················· | ···················· | ···················· | ···················· | ···················· | |
| T | 7519 hCoV-19/Australia/VIC16941/2020\|EPI_ISL_663651\|202 | ···················· | ···················· | ···················· | ···················· | ···················· | ···················· | |
| T | 7520 hCoV-19/Australia/VIC16942/2020\|EPI_ISL_663652\|202 | ···················· | ···················· | ···················· | ···················· | ···················· | ···················· | |
| T | 7521 hCoV-19/Australia/VIC16943/2020\|EPI_ISL_663653\|202 | ···················· | ···················· | ···················· | ···················· | ···················· | ···················· | |
| T | 7523 hCoV-19/Australia/VIC16949/2020\|EPI_ISL_663659\|202 | ···················· | ···················· | ···················· | ···················· | ···················· | ···················· | |
| T | 7525 hCoV-19/Australia/VIC16960/2020\|EPI_ISL_663661\|202 | ···················· | ···················· | ···················· | ···················· | ···················· | ···················· | |
| T | 7526 hCoV-19/Australia/VIC16962/2020\|EPI_ISL_663662\|202 | ···················· | ···················· | ···················· | ···················· | ···················· | ···················· | |
| T | 7527 hCoV-19/Australia/VIC16965/2020\|EPI_ISL_663663\|202 | ···················· | ···················· | ···················· | ···················· | ···················· | ···················· | |
| T | 7528 hCoV-19/Australia/VIC16967/2020\|EPI_ISL_663664\|202 | ···················· | ···················· | ···················· | ···················· | ···················· | ···················· | |
| T | 7529 hCoV-19/England/CAMB-1B9937/2020\|EPI_ISL_664952\|20 | ···················· | ···················· | ···················· | ···················· | ···················· | ···················· | |
| T | 7530 hCoV-19/England/CAMB-1B9964/2020\|EPI_ISL_664953\|20 | ···················· | ···················· | ···················· | ···················· | ···················· | ···················· | |
| T | 7531 hCoV-19/England/CAMB-1B9973/2020\|EPI_ISL_664954\|20 | ···················· | ···················· | ···················· | ···················· | ···················· | ···················· | |
| T | 7532 hCoV-19/England/CAMB-1B9982/2020\|EPI_ISL_664955\|20 | ···················· | ···················· | ···················· | ···················· | ···················· | ···················· | |
| T | 7533 hCoV-19/England/CAMB-1B9991/2020\|EPI_ISL_664956\|20 | ···················· | ···················· | ···················· | ···················· | ···················· | ···················· | |
| T | 7534 hCoV-19/England/CAMB-1B99A0/2020\|EPI_ISL_664957\|20 | ···················· | ···················· | ···················· | ···················· | ···················· | ···················· | |
| T | 7535 hCoV-19/England/CAMB-1B9A07/2020\|EPI_ISL_664958\|20 | ···················· | ···················· | ···················· | ···················· | ···················· | ···················· | |
| T | 7536 hCoV-19/England/CAMB-1B9B6E/2020\|EPI_ISL_664959\|20 | ···················· | ···················· | ···················· | ···················· | ···················· | ···················· | |
| T | 7537 hCoV-19/England/CAMB-1B9B8C/2020\|EPI_ISL_664960\|20 | ···················· | ···················· | ···················· | ···················· | ···················· | ···················· | |
| T | 7538 hCoV-19/England/CAMB-1B9B9B/2020\|EPI_ISL_664961\|20 | ···················· | ···················· | ···················· | ···················· | ···················· | ···················· | |
| T | 7539 hCoV-19/England/CAMB-1B9BC8/2020\|EPI_ISL_664962\|20 | ···················· | ···················· | ···················· | ···················· | ···················· | ···················· | |
| T | 7540 hCoV-19/England/CAMB-1B9BF5/2020\|EPI_ISL_664963\|20 | ···················· | ···················· | ···················· | ···················· | ···················· | ···················· | |
| T | 7541 hCoV-19/England/CAMB-1B9C2F/2020\|EPI_ISL_664964\|20 | ···················· | ···················· | ···················· | ···················· | ···T················ | ···················· | |
| T | 7542 hCoV-19/England/CAMB-1B9C3E/2020\|EPI_ISL_664965\|20 | ···················· | ···················· | ···················· | ···················· | ···················· | ···················· | |
| T | 7543 hCoV-19/England/CAMB-1B9C4D/2020\|EPI_ISL_664966\|20 | ···················· | ···················· | ···················· | ···················· | ···················· | ···················· | |
| T | 7544 hCoV-19/England/CAMB-1B9C5C/2020\|EPI_ISL_664967\|20 | ···················· | ···················· | ···················· | ···················· | ···················· | ···················· | |
| T | 7545 hCoV-19/England/CAMB-1B9CA7/2020\|EPI_ISL_664968\|20 | ···················· | ···················· | ···················· | ···················· | ···················· | ···················· | |
| T | 7546 hCoV-19/England/CAMB-1BA0B9/2020\|EPI_ISL_664969\|20 | ···················· | ···················· | ···················· | ···················· | ·····T·············· | ···················· | |
| T | 7547 hCoV-19/England/CAMB-1BA110/2020\|EPI_ISL_664970\|20 | ···················· | ···················· | ···················· | ···················· | ·····T·············· | ···················· | |
| T | 7548 hCoV-19/England/CAMB-1BA15C/2020\|EPI_ISL_664971\|20 | ···················· | ···················· | ···················· | ···················· | ···················· | ···················· | |
| T | 7549 hCoV-19/England/CAMB-1BA17A/2020\|EPI_ISL_664972\|20 | ···················· | ···················· | ···················· | ···················· | ···················· | ···················· | |
| T | 7550 hCoV-19/England/CAMB-1BA189/2020\|EPI_ISL_664973\|20 | ···················· | ···················· | ···················· | ···················· | ···················· | ···················· | |
| T | 7551 hCoV-19/England/CAMB-1BA1A7/2020\|EPI_ISL_664974\|20 | ···················· | ···················· | ···················· | ···················· | ···················· | ···················· | |
| T | 7552 hCoV-19/England/CAMB-1BA24A/2020\|EPI_ISL_664975\|20 | ···················· | ···················· | ···················· | ···················· | ···················· | ···················· | |
| T | 7553 hCoV-19/England/CAMB-1BA30B/2020\|EPI_ISL_664976\|20 | ···················· | ···················· | ···················· | ···················· | ···················· | ···················· | |
| T | 7554 hCoV-19/England/CAMB-1BA347/2020\|EPI_ISL_664977\|20 | ···················· | ···················· | ···················· | ···················· | ···················· | ···················· | |
| T | 7555 hCoV-19/England/CAMB-1BA3CF/2020\|EPI_ISL_664978\|20 | ···················· | ···················· | ···················· | ···················· | ···················· | ···················· | |
| T | 7556 hCoV-19/England/CAMB-1BA3DE/2020\|EPI_ISL_664979\|20 | ···················· | ···················· | ···················· | ···················· | ···················· | ···················· | |
| T | 7557 hCoV-19/England/CAMB-1BA417/2020\|EPI_ISL_664980\|20 | ···················· | ···················· | ···················· | ···················· | ···················· | ···················· | |
| T | 7558 hCoV-19/England/CAMB-1BA426/2020\|EPI_ISL_664981\|20 | ···················· | ···················· | ···················· | ···················· | ···················· | ···················· | |
| T | 7559 hCoV-19/England/CAMB-1BA444/2020\|EPI_ISL_664982\|20 | ···················· | ···················· | ···················· | ···················· | ···T················ | ···················· | |
| T | 7560 hCoV-19/England/CAMB-1BA49F/2020\|EPI_ISL_664983\|20 | ···················· | ···················· | ················G··· | ···················· | ·····T·············· | ···················· | |

| | | | | | | | |
|---|---|---|---|---|---|---|---|
| T | 7561 hCoV-19/England/CAMB-1BA4BD/2020\|EPI_ISL_664984\|20 | ···················· | ···················· | ···················· | ···················· | ···················· | ··A················· |
| T | 7562 hCoV-19/England/CAMB-1BA5AB/2020\|EPI_ISL_664989\|20 | ···················· | ···················· | ···················· | ···················· | ···················· | ···················· |
| T | 7563 hCoV-19/England/CAMB-1BA620/2020\|EPI_ISL_664990\|20 | ···················· | ···················· | ···················· | ···················· | ···················· | ···················· |
| T | 7564 hCoV-19/England/CAMB-1BA63F/2020\|EPI_ISL_664991\|20 | ···················· | ···················· | ···················· | ···················· | ···················· | ···················· |
| T | 7565 hCoV-19/England/CAMB-1BA699/2020\|EPI_ISL_664992\|20 | ···················· | ···················· | ···················· | ···················· | ·····T·············· | ···················· |
| T | 7566 hCoV-19/England/CAMB-1BA6C6/2020\|EPI_ISL_664993\|20 | ···················· | ···················· | ···················· | ···················· | ···················· | ···················· |
| T | 7567 hCoV-19/England/CAMB-1BA72D/2020\|EPI_ISL_664994\|20 | ···················· | ···················· | ···················· | ···················· | ···················· | ···················· |
| T | 7568 hCoV-19/England/CAMB-1BA769/2020\|EPI_ISL_664995\|20 | ···················· | ···················· | ···················· | ···················· | ···················· | ···················· |
| T | 7569 hCoV-19/USA/OR-OHSU-2039/2020\|EPI_ISL_667100\|2020- | ···················· | ·A·················· | ···················· | ···················· | ···················· | ···················· |
| T | 7570 hCoV-19/USA/OR-OHSU-2043/2020\|EPI_ISL_667101\|2020- | ···················· | ·A·················· | ···················· | ···················· | ···················· | ···················· |
| T | 7571 hCoV-19/USA/WA-OHSU-2045/2020\|EPI_ISL_667102\|2020- | ···················· | ·A·················· | ···················· | ···················· | ···················· | ···················· |
| T | 7572 hCoV-19/USA/OR-OHSU-2055/2020\|EPI_ISL_667104\|2020- | ···················· | ···················· | ···················· | ···················· | ···················· | ···················· |
| T | 7573 hCoV-19/USA/WA-OHSU-2057/2020\|EPI_ISL_667105\|2020- | ···················· | ···················· | ···················· | ···················· | ···················· | ···················· |
| T | 7574 hCoV-19/USA/OR-OHSU-2067/2020\|EPI_ISL_667106\|2020- | ···················· | ···················· | ···················· | ···················· | ···················· | ···················· |
| T | 7575 hCoV-19/USA/OR-OHSU-2069/2020\|EPI_ISL_667107\|2020- | ···················· | ···················· | ···················· | ···················· | ···················· | ···················· |
| T | 7576 hCoV-19/USA/OR-OHSU-2073/2020\|EPI_ISL_667109\|2020- | ···················· | ···················· | ···················· | ···················· | ···················· | ···················· |
| T | 7577 hCoV-19/USA/OR-OHSU-2075/2020\|EPI_ISL_667110\|2020- | ···················· | ···················· | ···················· | ···················· | ···················· | ···················· |
| T | 7578 hCoV-19/USA/OR-OHSU-2083/2020\|EPI_ISL_667112\|2020- | ···················· | ·A·················· | ···················· | ···················· | ···················· | ···················· |
| T | 7579 hCoV-19/USA/OR-OHSU-2087/2020\|EPI_ISL_667113\|2020- | ···················· | ···················· | ···················· | ···················· | ···················· | ···················· |
| T | 7580 hCoV-19/USA/OR-OHSU-2093/2020\|EPI_ISL_667114\|2020- | ···················· | ···················· | ···················· | ···················· | ···················· | ···················· |
| T | 7581 hCoV-19/USA/OR-OHSU-2103/2020\|EPI_ISL_667116\|2020- | ···················· | ···················· | ···················· | ···················· | ···················· | ···················· |
| T | 7582 hCoV-19/USA/OR-OHSU-2105/2020\|EPI_ISL_667117\|2020- | ···················· | ···················· | ···················· | ···················· | ···················· | ···················· |
| T | 7583 hCoV-19/USA/OR-OHSU-2119/2020\|EPI_ISL_667119\|2020- | ···················· | ···················· | ···················· | ···················· | ···················· | ···················· |
| T | 7584 hCoV-19/USA/OR-OHSU-2123/2020\|EPI_ISL_667121\|2020- | ···················· | ···················· | ···················· | ···················· | ···················· | ···················· |
| T | 7585 hCoV-19/USA/OR-OHSU-2129/2020\|EPI_ISL_667122\|2020- | ···················· | ···················· | ···················· | ···················· | ···················· | ···················· |
| T | 7586 hCoV-19/USA/OR-OHSU-2131/2020\|EPI_ISL_667123\|2020- | ···················· | ···················· | ···················· | ···················· | ···················· | ···················· |
| T | 7587 hCoV-19/USA/OR-OHSU-2133/2020\|EPI_ISL_667124\|2020- | ···················· | ···················· | ···················· | ···················· | ···················· | ···················· |
| T | 7588 hCoV-19/USA/OR-OHSU-2135/2020\|EPI_ISL_667125\|2020- | ···················· | ·A·················· | ···················· | ···················· | ···················· | ···················· |
| T | 7589 hCoV-19/USA/OR-OHSU-2143/2020\|EPI_ISL_667126\|2020- | ···················· | ···················· | ···················· | ···················· | ···················· | ···················· |
| T | 7590 hCoV-19/USA/OR-OHSU-2145/2020\|EPI_ISL_667127\|2020- | ···················· | ·A·················· | ···················· | ···················· | ···················· | ···················· |
| T | 7591 hCoV-19/USA/OR-OHSU-2151/2020\|EPI_ISL_667128\|2020- | ···················· | ···················· | ···················· | ···················· | ···················· | ···················· |
| T | 7592 hCoV-19/USA/OR-OHSU-2153/2020\|EPI_ISL_667129\|2020- | ···················· | ···················· | ···················· | ···················· | ···················· | ···················· |
| T | 7593 hCoV-19/USA/OR-OHSU-2159/2020\|EPI_ISL_667130\|2020- | ···················· | ···················· | ···················· | ···················· | ···················· | ···················· |
| T | 7594 hCoV-19/USA/OR-OHSU-2165/2020\|EPI_ISL_667131\|2020- | ···················· | ·A·················· | ···················· | ···················· | ···················· | ···················· |
| T | 7595 hCoV-19/USA/OR-OHSU-2167/2020\|EPI_ISL_667132\|2020- | ···················· | ·A·················· | ···················· | ···················· | ···················· | ···················· |
| T | 7596 hCoV-19/USA/OR-OHSU-2169/2020\|EPI_ISL_667133\|2020- | ···················· | ···················· | ···················· | ···················· | ···················· | ···················· |
| T | 7597 hCoV-19/USA/OR-OHSU-2171/2020\|EPI_ISL_667134\|2020- | ···················· | ·A·················· | ···················· | ···················· | ···················· | ···················· |
| T | 7598 hCoV-19/USA/OR-OHSU-2175/2020\|EPI_ISL_667135\|2020- | ···················· | ·A·················· | ···················· | ···················· | ···················· | ···················· |
| T | 7599 hCoV-19/USA/WA-OHSU-2177/2020\|EPI_ISL_667136\|2020- | ···················· | ···················· | ···················· | ···················· | ···················· | ···················· |
| T | 7600 hCoV-19/USA/OR-OHSU-2179/2020\|EPI_ISL_667137\|2020- | ···················· | ·A·················· | ···················· | ···················· | ···················· | ···················· |
| T | 7601 hCoV-19/USA/OR-OHSU-2181/2020\|EPI_ISL_667138\|2020- | ···················· | ·A·················· | ···················· | ···················· | ···················· | ···················· |
| T | 7602 hCoV-19/USA/OR-OHSU-2183/2020\|EPI_ISL_667139\|2020- | ···················· | ···················· | ···················· | ···················· | ···················· | ···················· |
| T | 7603 hCoV-19/USA/OR-OHSU-2187/2020\|EPI_ISL_667140\|2020- | ···················· | ···················· | ···················· | ···················· | ···················· | ···················· |
| T | 7604 hCoV-19/USA/OR-OHSU-2193/2020\|EPI_ISL_667142\|2020- | ···················· | ·A·················· | ···················· | ···················· | ···················· | ···················· |
| T | 7605 hCoV-19/USA/OR-OHSU-2195/2020\|EPI_ISL_667143\|2020- | ···················· | ·A·················· | ···················· | ···················· | ···················· | ···················· |
| T | 7606 hCoV-19/USA/OR-OHSU-2197/2020\|EPI_ISL_667144\|2020- | ···················· | ···················· | ···················· | ···················· | ···················· | ···················· |
| T | 7607 hCoV-19/USA/WA-OHSU-2201/2020\|EPI_ISL_667145\|2020- | ···················· | ···················· | ···················· | ···················· | ···················· | ···················· |
| T | 7608 hCoV-19/USA/OR-OHSU-2203/2020\|EPI_ISL_667146\|2020- | ···················· | ·A·················· | ···················· | ···················· | ···················· | ···················· |
| T | 7609 hCoV-19/USA/OR-OHSU-2211/2020\|EPI_ISL_667147\|2020- | ···················· | ···················· | ···················· | ···················· | ···················· | ···················· |
| T | 7610 hCoV-19/USA/OR-OHSU-2215/2020\|EPI_ISL_667149\|2020- | ···················· | ·A·················· | ···················· | ···················· | ···················· | ···················· |
| T | 7611 hCoV-19/Norway/4880/2020\|EPI_ISL_668417\|2020-10-21 | ···················· | ···················· | ···················· | ···················· | ···················· | ···················· |
| T | 7612 hCoV-19/Norway/4884/2020\|EPI_ISL_668418\|2020-10-18 | ···················· | ···················· | ···················· | ···················· | ···················· | ···················· |
| T | 7613 hCoV-19/Norway/4886/2020\|EPI_ISL_668419\|2020-10-23 | ···················· | ···················· | ···················· | ···················· | ···················· | ···················· |
| T | 7614 hCoV-19/Norway/4906/2020\|EPI_ISL_668428\|2020-10-26 | ···················· | ···················· | ···················· | ···················· | ···················· | ···················· |
| T | 7615 hCoV-19/Norway/4907/2020\|EPI_ISL_668429\|2020-10-24 | ···················· | ···················· | ···················· | ···················· | ···················· | ···················· |
| T | 7616 hCoV-19/Norway/4916/2020\|EPI_ISL_668430\|2020-10-25 | ···················· | ···················· | ···················· | ···················· | ···················· | ···················· |
| T | 7617 hCoV-19/Norway/4917/2020\|EPI_ISL_668431\|2020-10-25 | ···················· | ···················· | ···················· | ···················· | ···················· | ···················· |
| T | 7618 hCoV-19/Norway/4919/2020\|EPI_ISL_668432\|2020-10-24 | ···················· | ···················· | ···················· | ···················· | ···················· | ···················· |
| T | 7619 hCoV-19/Norway/4922/2020\|EPI_ISL_668433\|2020-10-25 | ···················· | ···················· | ···················· | ···················· | ···················· | ···················· |
| T | 7620 hCoV-19/Norway/4913/2020\|EPI_ISL_668434\|2020-10-25 | ···················· | ···················· | ···················· | ···················· | ···················· | ···················· |
| T | 7621 hCoV-19/Norway/4937/2020\|EPI_ISL_668435\|2020-10-28 | ···················· | ···················· | ···················· | ···················· | ···················· | ···················· |
| T | 7622 hCoV-19/Norway/4948/2020\|EPI_ISL_668436\|2020-10-29 | ···················· | ·A·················· | ···················· | ···················· | ···················· | ··A················· |
| T | 7623 hCoV-19/Norway/4960/2020\|EPI_ISL_668437\|2020-10-29 | ···················· | ···················· | ···················· | ···················· | ···················· | ···················· |
| T | 7624 hCoV-19/Norway/3780/2020\|EPI_ISL_668438\|2020-08-30 | ···················· | ···················· | ···················· | ···················· | ···················· | ···················· |
| T | 7625 hCoV-19/Norway/3784/2020\|EPI_ISL_668439\|2020-09-29 | ···················· | ···················· | ···················· | ···················· | ···················· | ···················· |
| T | 7626 hCoV-19/Norway/3787/2020\|EPI_ISL_668440\|2020-09-02 | ···················· | ···················· | ···················· | ···················· | ···················· | ···················· |
| T | 7627 hCoV-19/Norway/4592/2020\|EPI_ISL_668441\|2020-10-15 | ···················· | ···················· | ···················· | ···················· | ···················· | ···················· |
| T | 7629 hCoV-19/Denmark/DCGC-10885/2020\|EPI_ISL_669700\|202 | ···················· | ···················· | ···················· | ···················· | ···················· | ···················· |
| T | 7630 hCoV-19/Denmark/DCGC-10895/2020\|EPI_ISL_669701\|202 | ···················· | ···················· | ···················· | ···················· | ···················· | ···················· |
| T | 7631 hCoV-19/Denmark/DCGC-10900/2020\|EPI_ISL_669702\|202 | ···················· | ···················· | ···················· | ···················· | ···················· | ···················· |
| T | 7632 hCoV-19/Denmark/DCGC-10903/2020\|EPI_ISL_669703\|202 | ···················· | ···················· | ···················· | ···················· | ···················· | ···················· |
| T | 7633 hCoV-19/Denmark/DCGC-10908/2020\|EPI_ISL_669704\|202 | ···················· | ···················· | ···················· | ···················· | ···················· | ···················· |
| T | 7634 hCoV-19/Denmark/DCGC-10912/2020\|EPI_ISL_669705\|202 | ···················· | ···················· | ···················· | ···················· | ···················· | ···················· |
| T | 7635 hCoV-19/Denmark/DCGC-10914/2020\|EPI_ISL_669706\|202 | ···················· | ···················· | ···················· | ···················· | ···················· | ···················· |
| T | 7636 hCoV-19/Denmark/DCGC-10924/2020\|EPI_ISL_669707\|202 | ···················· | ···················· | ···················· | ···················· | ···················· | ···················· |
| T | 7637 hCoV-19/Denmark/DCGC-10942/2020\|EPI_ISL_669708\|202 | ···················· | ···················· | ···················· | ···················· | ···················· | ···················· |
| T | 7638 hCoV-19/Denmark/DCGC-10946/2020\|EPI_ISL_669709\|202 | ···················· | ···················· | ···················· | ···················· | ···················· | ···················· |
| T | 7639 hCoV-19/Denmark/DCGC-10957/2020\|EPI_ISL_669710\|202 | ···················· | ···················· | ···················· | ···················· | ···················· | ···················· |
| T | 7640 hCoV-19/Denmark/DCGC-10960/2020\|EPI_ISL_669711\|202 | ···················· | ···················· | ···················· | ···················· | ···················· | ···················· |
| T | 7641 hCoV-19/Denmark/DCGC-11004/2020\|EPI_ISL_669712\|202 | ···················· | ···················· | ···················· | ···················· | ···················· | ···················· |

| | | | | | | | | |
|---|---|---|---|---|---|---|---|---|
| T | 7642 | hCoV-19/Denmark/DCGC-11009/2020\|EPI_ISL_669713\|202 | ···················· | ···················· | ····················· | ···················· | ···················· | ···················· |
| T | 7643 | hCoV-19/Denmark/DCGC-11025/2020\|EPI_ISL_669714\|202 | ···················· | ···················· | ····················· | ···················· | ···················· | ···················· |
| T | 7644 | hCoV-19/Denmark/DCGC-11044/2020\|EPI_ISL_669715\|202 | ···················· | ···················· | ····················· | ···················· | ···················· | ···················· |
| T | 7645 | hCoV-19/Denmark/DCGC-11080/2020\|EPI_ISL_669716\|202 | ···················· | ···················· | ····················· | ···················· | ···················· | ···················· |
| T | 7646 | hCoV-19/Denmark/DCGC-11099/2020\|EPI_ISL_669717\|202 | ···················· | ···················· | ····················· | ···················· | ···················· | ···················· |
| T | 7647 | hCoV-19/Denmark/DCGC-11107/2020\|EPI_ISL_669718\|202 | ···················· | ···················· | ····················· | ···················· | ···················· | ···················· |
| T | 7648 | hCoV-19/Denmark/DCGC-11113/2020\|EPI_ISL_669719\|202 | ···················· | ···················· | ····················· | ···················· | ···················· | ···················· |
| T | 7649 | hCoV-19/Denmark/DCGC-11115/2020\|EPI_ISL_669720\|202 | ···················· | ···················· | ····················· | ···················· | ···················· | ···················· |
| T | 7650 | hCoV-19/Denmark/DCGC-11120/2020\|EPI_ISL_669721\|202 | ···················· | ···················· | ····················· | ···················· | ···················· | ···················· |
| T | 7651 | hCoV-19/Denmark/DCGC-11123/2020\|EPI_ISL_669722\|202 | ···················· | ···················· | ····················· | ···················· | ···················· | ···················· |
| T | 7652 | hCoV-19/Denmark/DCGC-11132/2020\|EPI_ISL_669723\|202 | ···················· | ···················· | ····················· | ···················· | ···················· | ···················· |
| T | 7653 | hCoV-19/Denmark/DCGC-11160/2020\|EPI_ISL_669724\|202 | ···················· | ···················· | ····················· | ···················· | ···················· | ···················· |
| T | 7654 | hCoV-19/Denmark/DCGC-11161/2020\|EPI_ISL_669725\|202 | ···················· | ···················· | ····················· | ···················· | ···················· | ···················· |
| T | 7655 | hCoV-19/Denmark/DCGC-11102/2020\|EPI_ISL_669748\|202 | ···················· | ···················· | ····················· | ···················· | ···················· | ···················· |
| T | 7656 | hCoV-19/Denmark/DCGC-11104/2020\|EPI_ISL_669749\|202 | ···················· | ···················· | ····················· | ···················· | ···················· | ···················· |
| T | 7657 | hCoV-19/Denmark/DCGC-11129/2020\|EPI_ISL_669750\|202 | ···················· | ···················· | ····················· | ···················· | ···················· | ···················· |
| T | 7658 | hCoV-19/Denmark/DCGC-11424/2020\|EPI_ISL_670746\|202 | ···················· | ·N·················· | ····················· | ···················· | ···················· | ···················· |
| T | 7659 | hCoV-19/Denmark/DCGC-11498/2020\|EPI_ISL_670747\|202 | ···················· | ···················· | ····················· | ···················· | ···················· | ···················· |
| T | 7660 | hCoV-19/Denmark/DCGC-11212/2020\|EPI_ISL_670772\|202 | ···················· | ···················· | ····················· | ···················· | ···················· | ···················· |
| T | 7661 | hCoV-19/Denmark/DCGC-11215/2020\|EPI_ISL_670773\|202 | ···················· | ···················· | ····················· | ···················· | ···················· | ···················· |
| T | 7662 | hCoV-19/Denmark/DCGC-11219/2020\|EPI_ISL_670774\|202 | ···················· | ···················· | ····················· | ···················· | ···················· | ···················· |
| T | 7663 | hCoV-19/Denmark/DCGC-11220/2020\|EPI_ISL_670775\|202 | ···················· | ···················· | ····················· | ···················· | ···················· | ···················· |
| T | 7664 | hCoV-19/Denmark/DCGC-11222/2020\|EPI_ISL_670776\|202 | ···················· | ···················· | ····················· | ···················· | ···················· | ···················· |
| T | 7665 | hCoV-19/Denmark/DCGC-11224/2020\|EPI_ISL_670777\|202 | ···················· | ···················· | ····················· | ···················· | ···················· | ···················· |
| T | 7666 | hCoV-19/Denmark/DCGC-11227/2020\|EPI_ISL_670778\|202 | ···················· | ···················· | ····················· | ···················· | ···················· | ···················· |
| T | 7667 | hCoV-19/Denmark/DCGC-11228/2020\|EPI_ISL_670779\|202 | ···················· | ···················· | ····················· | ···················· | ···················· | ···················· |
| T | 7668 | hCoV-19/Denmark/DCGC-11230/2020\|EPI_ISL_670780\|202 | ···················· | ···················· | ····················· | ···················· | ···················· | ···················· |
| T | 7669 | hCoV-19/Denmark/DCGC-11231/2020\|EPI_ISL_670781\|202 | ···················· | ···················· | ····················· | ···················· | ···················· | ···················· |
| T | 7670 | hCoV-19/Denmark/DCGC-11232/2020\|EPI_ISL_670782\|202 | ···················· | ···················· | ····················· | ···················· | ···················· | ···················· |
| T | 7671 | hCoV-19/Denmark/DCGC-11233/2020\|EPI_ISL_670783\|202 | ···················· | ···················· | ····················· | ···················· | ···················· | ···················· |
| T | 7672 | hCoV-19/Denmark/DCGC-11234/2020\|EPI_ISL_670784\|202 | ···················· | A··················· | ····················· | ···················· | ···················· | ···················· |
| T | 7673 | hCoV-19/Denmark/DCGC-11235/2020\|EPI_ISL_670785\|202 | ···················· | ···················· | ····················· | ···················· | ···················· | ···················· |
| T | 7674 | hCoV-19/Denmark/DCGC-11236/2020\|EPI_ISL_670786\|202 | ···················· | ···················· | ····················· | ···················· | ···················· | ···················· |
| T | 7675 | hCoV-19/Denmark/DCGC-11237/2020\|EPI_ISL_670787\|202 | ···················· | ···················· | ····················· | ···················· | ···················· | ···················· |
| T | 7676 | hCoV-19/Denmark/DCGC-11238/2020\|EPI_ISL_670788\|202 | ···················· | ···················· | ····················· | ···················· | ···················· | ···················· |
| T | 7677 | hCoV-19/Denmark/DCGC-11240/2020\|EPI_ISL_670789\|202 | ···················· | ···················· | ····················· | ···················· | ···················· | ···················· |
| T | 7678 | hCoV-19/Denmark/DCGC-11241/2020\|EPI_ISL_670790\|202 | ···················· | ···················· | ····················· | ···················· | ···················· | ···················· |
| T | 7679 | hCoV-19/Denmark/DCGC-11244/2020\|EPI_ISL_670791\|202 | ···················· | ···················· | ····················· | ···················· | ···················· | ···················· |
| T | 7680 | hCoV-19/Denmark/DCGC-11247/2020\|EPI_ISL_670792\|202 | ···················· | ·A·················· | ····················· | ···················· | ···················· | ···················· |
| T | 7681 | hCoV-19/Norway/4764/2020\|EPI_ISL_668400\|2020-10-22 | ···················· | ···················· | ····················· | ···················· | ···················· | ···················· |
| T | 7682 | hCoV-19/Norway/4779/2020\|EPI_ISL_668401\|2020-10-19 | ···················· | ···················· | ····················· | ···················· | ···················· | ···················· |
| T | 7683 | hCoV-19/Norway/4781/2020\|EPI_ISL_668402\|2020-10-15 | ···················· | ···················· | ····················· | ···················· | ···················· | ···················· |
| T | 7684 | hCoV-19/Norway/4460/2020\|EPI_ISL_668405\|2020-10-12 | ···················· | ·A·················· | ····················· | ···················· | ···················· | ··A················· |
| T | 7685 | hCoV-19/Norway/4535/2020\|EPI_ISL_668406\|2020-10-06 | ···················· | ···················· | ····················· | ···················· | ···················· | ···················· |
| T | 7686 | hCoV-19/Norway/4795/2020\|EPI_ISL_668408\|2020-10-22 | ···················· | ···················· | ····················· | ···················· | ···················· | ···················· |
| T | 7687 | hCoV-19/Norway/4797/2020\|EPI_ISL_668409\|2020-10-23 | ···················· | ···················· | ····················· | ···················· | ···················· | ···················· |
| T | 7688 | hCoV-19/Norway/4810/2020\|EPI_ISL_668410\|2020-10-26 | ···················· | ·A·················· | ····················· | ···················· | ···················· | ··A················· |
| T | 7689 | hCoV-19/Norway/4811/2020\|EPI_ISL_668411\|2020-10-26 | ···················· | ·A·················· | ····················· | ···················· | ···················· | ··A················· |
| T | 7690 | hCoV-19/Norway/4856/2020\|EPI_ISL_668412\|2020-10-20 | ···················· | ···················· | ····················· | ···················· | ···················· | ···················· |
| T | 7691 | hCoV-19/Norway/4858/2020\|EPI_ISL_668413\|2020-10-23 | ···················· | ···················· | ····················· | ···················· | ···················· | ···················· |
| T | 7692 | hCoV-19/Norway/4865/2020\|EPI_ISL_668415\|2020-10-23 | ···················· | ···················· | ····················· | ···················· | ···················· | ···················· |
| T | 7693 | hCoV-19/Norway/4868/2020\|EPI_ISL_668416\|2020-10-20 | ···················· | ···················· | ····················· | ···················· | ···················· | ···················· |
| T | 7694 | hCoV-19/Norway/4885/2020\|EPI_ISL_668420\|2020-10-20 | ···················· | ···················· | ····················· | ···················· | ···················· | ···················· |
| T | 7695 | hCoV-19/Norway/4889/2020\|EPI_ISL_668421\|2020-10-21 | ···················· | ···················· | ····················· | ···················· | ···················· | ···················· |
| T | 7696 | hCoV-19/Norway/4891/2020\|EPI_ISL_668422\|2020-10-22 | ···················· | ···················· | ····················· | ···················· | ···················· | ···················· |
| T | 7697 | hCoV-19/Norway/4893/2020\|EPI_ISL_668423\|2020-10-23 | ···················· | ···················· | ····················· | ···················· | ···················· | ···················· |
| T | 7698 | hCoV-19/Norway/4896/2020\|EPI_ISL_668424\|2020-10-26 | ···················· | ···················· | ····················· | ···················· | ···················· | ···················· |
| T | 7699 | hCoV-19/Norway/4897/2020\|EPI_ISL_668425\|2020-10-25 | ···················· | ···················· | ····················· | ···················· | ···················· | ···················· |
| T | 7700 | hCoV-19/Norway/4898/2020\|EPI_ISL_668426\|2020-10-25 | ···················· | ·A·················· | ····················· | ···················· | ···················· | ···················· |
| T | 7701 | hCoV-19/Norway/4903/2020\|EPI_ISL_668427\|2020-10-25 | ···················· | ···················· | ····················· | ···················· | ···················· | ···················· |
| T | 7702 | hCoV-19/Denmark/DCGC-11180/2020\|EPI_ISL_669726\|202 | ···················· | ···················· | ····················· | ···················· | ···················· | ···················· |
| T | 7703 | hCoV-19/Denmark/DCGC-11182/2020\|EPI_ISL_669727\|202 | ···················· | ···················· | ····················· | ···················· | ···················· | ···················· |
| T | 7704 | hCoV-19/Denmark/DCGC-11201/2020\|EPI_ISL_669728\|202 | ···················· | ···················· | ····················· | ···················· | ···················· | ···················· |
| T | 7705 | hCoV-19/Denmark/DCGC-11203/2020\|EPI_ISL_669729\|202 | ···················· | ···················· | ····················· | ···················· | ···················· | ···················· |
| T | 7706 | hCoV-19/Denmark/DCGC-11207/2020\|EPI_ISL_669730\|202 | ···················· | ···················· | ····················· | ···················· | ···················· | ···················· |
| T | 7707 | hCoV-19/Denmark/DCGC-10255/2020\|EPI_ISL_669733\|202 | ···················· | ···················· | ····················· | ···················· | ···················· | ···················· |
| T | 7708 | hCoV-19/Denmark/DCGC-10263/2020\|EPI_ISL_669734\|202 | ···················· | ·A·················· | ····················· | ···················· | ···················· | ···················· |
| T | 7709 | hCoV-19/Denmark/DCGC-10267/2020\|EPI_ISL_669735\|202 | ···················· | ···················· | ····················· | ···················· | ···················· | ···················· |
| T | 7710 | hCoV-19/Denmark/DCGC-10474/2020\|EPI_ISL_669736\|202 | ···················· | ·A·················· | ····················· | ···················· | ···················· | ··A················· |
| T | 7711 | hCoV-19/Denmark/DCGC-10552/2020\|EPI_ISL_669737\|202 | ···················· | ···················· | ····················· | ···················· | ···················· | ···················· |
| T | 7712 | hCoV-19/Denmark/DCGC-10640/2020\|EPI_ISL_669738\|202 | ···················· | ···················· | ····················· | ···················· | ···················· | ···················· |
| T | 7713 | hCoV-19/Denmark/DCGC-10657/2020\|EPI_ISL_669739\|202 | ···················· | ···················· | ····················· | ···················· | ···················· | ···················· |
| T | 7714 | hCoV-19/Denmark/DCGC-10800/2020\|EPI_ISL_669740\|202 | ···················· | ···················· | ····················· | ···················· | ···················· | ···················· |
| T | 7715 | hCoV-19/Denmark/DCGC-10862/2020\|EPI_ISL_669741\|202 | ···················· | ···················· | ····················· | ···················· | ···················· | ···················· |
| T | 7716 | hCoV-19/Denmark/DCGC-10886/2020\|EPI_ISL_669742\|202 | ···················· | ···················· | ····················· | ···················· | ···················· | ···················· |
| T | 7717 | hCoV-19/Denmark/DCGC-10959/2020\|EPI_ISL_669743\|202 | ···················· | ·A·················· | ····················· | ···················· | ···················· | ··A················· |
| T | 7718 | hCoV-19/Denmark/DCGC-10974/2020\|EPI_ISL_669744\|202 | ···················· | ···················· | ····················· | ···················· | ···················· | ···················· |
| T | 7719 | hCoV-19/Denmark/DCGC-11016/2020\|EPI_ISL_669745\|202 | ···················· | ···················· | ····················· | ···················· | ···················· | ···················· |
| T | 7720 | hCoV-19/Denmark/DCGC-11053/2020\|EPI_ISL_669746\|202 | ···················· | ·A·················· | ····················· | ···················· | ···················· | ··A················· |
| T | 7721 | hCoV-19/Denmark/DCGC-11066/2020\|EPI_ISL_669747\|202 | ···················· | ·A·················· | ····················· | ···················· | ···················· | ··A················· |

| | | | | | | | |
|---|---|---|---|---|---|---|---|
| T | 7722 hCoV-19/Denmark/DCGC-10723/2020\|EPI_ISL_669754\|202 | .................... | .................... | .................... | .................... | .................... | .................... |
| T | 7723 hCoV-19/Denmark/DCGC-11083/2020\|EPI_ISL_669755\|202 | .................... | .................... | .................... | .................... | .................... | .................... |

Group 6 - Dec 2020

| A | B | C | D | E | F | G | H | I | J | K | L |
|---|---|---|---|---|---|---|---|---|---|---|---|
| Database | Database ID | GISAID ID | GCTGCAATCGTGCTACAACT | TGAACTGTTGCGACTACGTG | TTCGGAAGAGACAGGTACGTT | CACACAATCGAAGCGCAGTA | GCTGGTGCTGCAGCTTATTA | AGGGTCAAGTGCACAGTCTA | mismatches? | mismatches? | primer set? |
| U | 1 | hCoV-19/England/CAMC-B98372/2020\|EPI_ISL_673701\|20 | ···················· | ···················· | ···················· | ···················· | ···················· | ···················· | | | |
| U | 2 | hCoV-19/England/MILK-B94B7E/2020\|EPI_ISL_673702\|20 | ···················· | ···················· | ···················· | ···················· | ···················· | ···················· | | | |
| U | 3 | hCoV-19/England/CAMC-B9898F/2020\|EPI_ISL_673704\|20 | ···················· | ···················· | ···················· | ···················· | ···················· | ···················· | | | |
| U | 4 | hCoV-19/England/CAMC-B98E35/2020\|EPI_ISL_673706\|20 | ···················· | ···················· | ···················· | ···················· | ···················· | ···················· | | | |
| U | 5 | hCoV-19/England/CAMC-B9AE8E/2020\|EPI_ISL_673707\|20 | ···················· | ···················· | ···················· | ···················· | ···················· | ···················· | | | |
| U | 6 | hCoV-19/England/MILK-B954EB/2020\|EPI_ISL_673708\|20 | ···················· | ·A·················· | ···················· | ···················· | ···················· | ···················· | | | |
| U | 7 | hCoV-19/England/MILK-B9520F/2020\|EPI_ISL_673709\|20 | ···················· | ···················· | ···················· | ···················· | ···················· | ···················· | | | |
| U | 8 | hCoV-19/England/CAMC-B99E61/2020\|EPI_ISL_673710\|20 | ···················· | ···················· | ···················· | ···················· | ···················· | ···················· | | | |
| U | 9 | hCoV-19/England/CAMC-B98D38/2020\|EPI_ISL_673711\|20 | ···················· | ···················· | ···················· | ···················· | ···················· | ···················· | | | |
| U | 10 | hCoV-19/England/CAMC-B98C59/2020\|EPI_ISL_673713\|20 | ···················· | ···················· | ···················· | ···················· | ···················· | ···················· | | | |
| U | 11 | hCoV-19/England/CAMC-B98E08/2020\|EPI_ISL_673714\|20 | ···················· | ···················· | ···················· | ···················· | ···················· | ···················· | | | |
| U | 12 | hCoV-19/England/MILK-B9515D/2020\|EPI_ISL_673715\|20 | ···················· | ···················· | ···················· | ···················· | ···················· | ···················· | | | |
| U | 13 | hCoV-19/England/CAMC-B9A60E/2020\|EPI_ISL_673716\|20 | ···················· | ·A·················· | ···················· | ···················· | ···················· | ··A················· | | | |
| U | 14 | hCoV-19/England/CAMC-B994AB/2020\|EPI_ISL_673717\|20 | ···················· | ···················· | ···················· | ···················· | ···················· | ···················· | | | |
| U | 15 | hCoV-19/England/CAMC-B9A219/2020\|EPI_ISL_673718\|20 | ···················· | ···················· | ···················· | ···················· | ···················· | ···················· | | | |
| U | 16 | hCoV-19/England/CAMC-B9A307/2020\|EPI_ISL_673719\|20 | ···················· | ···················· | ···················· | ···················· | ·····T·············· | ···················· | | | |
| U | 17 | hCoV-19/England/CAMC-B999E8/2020\|EPI_ISL_673720\|20 | ···················· | ···················· | ···················· | ···················· | ···················· | ···················· | | | |
| U | 18 | hCoV-19/England/CAMC-B98DCF/2020\|EPI_ISL_673721\|20 | ···················· | ···················· | ···················· | ···················· | ···················· | ···················· | | | |
| U | 19 | hCoV-19/England/CAMC-B989BC/2020\|EPI_ISL_673725\|20 | ···················· | ···················· | ···················· | ···················· | ···················· | ···················· | | | |
| U | 20 | hCoV-19/England/MILK-B94C99/2020\|EPI_ISL_673726\|20 | ···················· | ···················· | ···················· | ···················· | ···················· | ···················· | | | |
| U | 21 | hCoV-19/England/MILK-B952D2/2020\|EPI_ISL_673729\|20 | ···················· | ···················· | ···················· | ···················· | ·····T·············· | ···················· | | | |
| U | 22 | hCoV-19/England/MILK-B950F6/2020\|EPI_ISL_673730\|20 | ···················· | ···················· | ···················· | ···················· | ···················· | ···················· | | | |
| U | 23 | hCoV-19/England/CAMC-B989DA/2020\|EPI_ISL_673731\|20 | ···················· | ···················· | ···················· | ···················· | ···················· | ···················· | | | |
| U | 24 | hCoV-19/England/CAMC-B98327/2020\|EPI_ISL_673732\|20 | ···················· | ···················· | ···················· | ···················· | ···················· | ···················· | | | |
| U | 25 | hCoV-19/England/MILK-B951E4/2020\|EPI_ISL_673733\|20 | ···················· | ···················· | ···················· | ···················· | ···················· | ···················· | | | |
| U | 26 | hCoV-19/England/CAMC-B998EB/2020\|EPI_ISL_673734\|20 | ···················· | ···················· | ···················· | ···················· | ···················· | ···················· | | | |
| U | 27 | hCoV-19/England/CAMC-B9A668/2020\|EPI_ISL_673736\|20 | ···················· | ···················· | ···················· | ···················· | ·····T·············· | ···················· | | | |
| U | 28 | hCoV-19/England/CAMC-B9997F/2020\|EPI_ISL_673739\|20 | ···················· | ···················· | ···················· | ···················· | ·····T·············· | ···················· | | | |
| U | 29 | hCoV-19/England/CAMC-B9872B/2020\|EPI_ISL_673741\|20 | ···················· | ···················· | ···················· | ···················· | ···················· | ···················· | | | |
| U | 30 | hCoV-19/England/CAMC-B98D0B/2020\|EPI_ISL_673743\|20 | ···················· | ···················· | ···················· | ···················· | ···················· | ···················· | | | |
| U | 31 | hCoV-19/England/CAMC-B989F8/2020\|EPI_ISL_673744\|20 | ···················· | ···················· | ···················· | ···················· | ···················· | ···················· | | | |
| U | 32 | hCoV-19/England/MILK-B95111/2020\|EPI_ISL_673745\|20 | ···················· | ·A·················· | ···················· | ···················· | ···················· | ··A················· | | | |
| U | 33 | hCoV-19/England/CAMC-B9903E/2020\|EPI_ISL_673747\|20 | ···················· | ·A·················· | ···················· | ···················· | ···················· | ··A················· | | | |
| U | 34 | hCoV-19/England/MILK-B94BF6/2020\|EPI_ISL_673750\|20 | ···················· | ···················· | ···················· | ···················· | ···················· | ···················· | | | |
| U | 35 | hCoV-19/England/CAMC-B98A40/2020\|EPI_ISL_673751\|20 | ···················· | ···················· | ···················· | ···················· | ···················· | ···················· | | | |
| U | 36 | hCoV-19/England/CAMC-B9999D/2020\|EPI_ISL_673753\|20 | ···················· | ···················· | ···················· | ···················· | ···················· | ···················· | | | |
| U | 37 | hCoV-19/England/CAMC-B98891/2020\|EPI_ISL_673755\|20 | ···················· | ···················· | ···················· | ···················· | ···················· | ···················· | | | |
| U | 38 | hCoV-19/England/CAMC-B983EB/2020\|EPI_ISL_673756\|20 | ···················· | ···················· | ···················· | ···················· | ···················· | ···················· | | | |
| U | 39 | hCoV-19/England/CAMC-B98512/2020\|EPI_ISL_673757\|20 | ···················· | ···················· | ···················· | ···················· | ···················· | ···················· | | | |
| U | 40 | hCoV-19/England/CAMC-B9AE33/2020\|EPI_ISL_673758\|20 | ···················· | ···················· | ···················· | ···················· | ···················· | ···················· | | | |
| U | 41 | hCoV-19/England/CAMC-B9A57A/2020\|EPI_ISL_673759\|20 | ···················· | ···················· | ···················· | ···················· | ···················· | ···················· | | | |
| U | 42 | hCoV-19/England/MILK-B9580D/2020\|EPI_ISL_673760\|20 | ···················· | ···················· | ···················· | ···················· | ···················· | ···················· | | | |
| U | 43 | hCoV-19/England/CAMC-B985B8/2020\|EPI_ISL_673761\|20 | ···················· | ···················· | ···················· | ···················· | ·····T·············· | ···················· | | | |
| U | 44 | hCoV-19/England/CAMC-B999D9/2020\|EPI_ISL_673762\|20 | ···················· | ···················· | ···················· | ···················· | ···················· | ···················· | | | |
| U | 45 | hCoV-19/England/CAMC-B9A228/2020\|EPI_ISL_673763\|20 | ···················· | ···················· | ···················· | ···················· | ···················· | ···················· | | | |
| U | 46 | hCoV-19/England/MILK-B94FEB/2020\|EPI_ISL_673765\|20 | ···················· | ···················· | ···················· | ···················· | ···················· | ···················· | | | |
| U | 47 | hCoV-19/England/MILK-B95A61/2020\|EPI_ISL_673767\|20 | ···················· | ···················· | ···················· | ···················· | ·····T·············· | ···················· | | | |
| U | 48 | hCoV-19/England/CAMC-B994C9/2020\|EPI_ISL_673769\|20 | ···················· | ···················· | ···················· | ···················· | ···················· | ···················· | | | |
| U | 49 | hCoV-19/England/CAMC-B987A3/2020\|EPI_ISL_673772\|20 | ···················· | ···················· | ···················· | ···················· | ···················· | ···················· | | | |
| U | 50 | hCoV-19/England/CAMC-B9A5E3/2020\|EPI_ISL_673773\|20 | ···················· | ···················· | ···················· | ···················· | ···················· | ···················· | | | |
| U | 51 | hCoV-19/England/CAMC-B98CC2/2020\|EPI_ISL_673774\|20 | ···················· | ···················· | ···················· | ···················· | ···················· | ···················· | | | |
| U | 52 | hCoV-19/England/CAMC-B98749/2020\|EPI_ISL_673775\|20 | ···················· | ···················· | ···················· | ···················· | ···················· | ···················· | | | |
| U | 53 | hCoV-19/England/CAMC-B98C77/2020\|EPI_ISL_673776\|20 | ···················· | ···················· | ···················· | ···················· | ·····T·············· | ···················· | | | |
| U | 54 | hCoV-19/England/CAMC-B988BF/2020\|EPI_ISL_673777\|20 | ···················· | ···················· | ···················· | ···················· | ·····T·············· | ···················· | | | |
| U | 55 | hCoV-19/England/CAMC-B987D0/2020\|EPI_ISL_673779\|20 | ···················· | ···················· | ···················· | ···················· | ···················· | ···················· | | | |
| U | 56 | hCoV-19/England/CAMC-B98A31/2020\|EPI_ISL_673782\|20 | ···················· | ···················· | ···················· | ···················· | ·····T·············· | ···················· | | | |
| U | 57 | hCoV-19/England/CAMC-B9948D/2020\|EPI_ISL_673784\|20 | ···················· | ···················· | ···················· | ···················· | ···················· | ···················· | | | |
| U | 58 | hCoV-19/England/MILK-B95515/2020\|EPI_ISL_673785\|20 | ···················· | ···················· | ···················· | ···················· | ···················· | ···················· | | | |
| U | 59 | hCoV-19/England/MILK-B95102/2020\|EPI_ISL_673787\|20 | ···················· | ···················· | ···················· | ···················· | ···················· | ···················· | | | |
| U | 60 | hCoV-19/England/CAMC-B98794/2020\|EPI_ISL_673788\|20 | ···················· | ···················· | ···················· | ···················· | ···················· | ···················· | | | |
| U | 61 | hCoV-19/England/CAMC-B98873/2020\|EPI_ISL_673789\|20 | ···················· | ···················· | ···················· | ···················· | ···················· | ···················· | | | |
| U | 62 | hCoV-19/England/MILK-B949DE/2020\|EPI_ISL_673790\|20 | ···················· | ···················· | ···················· | ···················· | ···················· | ···················· | | | |
| U | 63 | hCoV-19/England/CAMC-B9949C/2020\|EPI_ISL_673791\|20 | ···················· | ···················· | ···················· | ···················· | ···················· | ···················· | | | |
| U | 64 | hCoV-19/England/CAMC-B9A5C5/2020\|EPI_ISL_673795\|20 | ···················· | ·A·················· | ···················· | ···················· | ···················· | ··A················· | | | |
| U | 65 | hCoV-19/England/MILK-B952A5/2020\|EPI_ISL_673796\|20 | ···················· | ···················· | ···················· | ···················· | ···················· | ···················· | | | |
| U | 66 | hCoV-19/England/MILK-B95524/2020\|EPI_ISL_673798\|20 | ···················· | ···················· | ···················· | ···················· | ···················· | ···················· | | | |
| U | 67 | hCoV-19/USA/VA-DCLS-2171/2020\|EPI_ISL_677200\|2020- | ···················· | ···················· | ···················· | ···················· | ···················· | ···················· | | | |
| U | 68 | hCoV-19/USA/VA-DCLS-2173/2020\|EPI_ISL_677201\|2020- | ···················· | ···················· | ···················· | ···················· | ···················· | ···················· | | | |
| U | 69 | hCoV-19/USA/VA-DCLS-2174/2020\|EPI_ISL_677202\|2020- | ···················· | ···················· | ···················· | ···················· | ···················· | ···················· | | | |

| | | | | | | | | |
|---|---|---|---|---|---|---|---|---|
| U | 70 | hCoV-19/USA/VA-DCLS-2176/2020\|EPI_ISL_677203\|2020- | ···················· | ···················· | ····················· | ···················· | ···················· | ···················· |
| U | 71 | hCoV-19/USA/VA-DCLS-2177/2020\|EPI_ISL_677204\|2020- | ···················· | ···················· | ····················· | ···················· | ···················· | ···················· |
| U | 72 | hCoV-19/USA/VA-DCLS-2178/2020\|EPI_ISL_677205\|2020- | ···················· | ···················· | ····················· | ···················· | ···················· | ···················· |
| U | 73 | hCoV-19/USA/VA-DCLS-2180/2020\|EPI_ISL_677206\|2020- | ···················· | ···················· | ····················· | ···················· | ···················· | ···················· |
| U | 74 | hCoV-19/USA/VA-DCLS-2181/2020\|EPI_ISL_677207\|2020- | ···················· | ···················· | ····················· | ···················· | ···················· | ···················· |
| U | 75 | hCoV-19/USA/VA-DCLS-2182/2020\|EPI_ISL_677208\|2020- | ···················· | ···················· | ····················· | ···················· | ···················· | ···················· |
| U | 76 | hCoV-19/USA/VA-DCLS-2183/2020\|EPI_ISL_677209\|2020- | ···················· | ···················· | ····················· | ···················· | ···················· | ···················· |
| U | 77 | hCoV-19/USA/VA-DCLS-2184/2020\|EPI_ISL_677210\|2020- | ···················· | ···················· | ····················· | ···················· | ···················· | ···················· |
| U | 78 | hCoV-19/USA/VA-DCLS-2185/2020\|EPI_ISL_677211\|2020- | ···················· | ···················· | ····················· | ···················· | ···················· | ···················· |
| U | 79 | hCoV-19/USA/VA-DCLS-2187/2020\|EPI_ISL_677212\|2020- | ···················· | ···················· | ····················· | ···················· | ···················· | ···················· |
| U | 80 | hCoV-19/USA/VA-DCLS-2188/2020\|EPI_ISL_677213\|2020- | ···················· | ·A·················· | ····················· | ···················· | ···················· | ···················· |
| U | 81 | hCoV-19/USA/VA-DCLS-2037/2020\|EPI_ISL_677215\|2020- | ···················· | ···················· | ····················· | ···················· | ···················· | ···················· |
| U | 82 | hCoV-19/USA/VA-DCLS-2038/2020\|EPI_ISL_677216\|2020- | ···················· | ···················· | ····················· | ···················· | ···················· | ···················· |
| U | 83 | hCoV-19/USA/VA-DCLS-2043/2020\|EPI_ISL_677217\|2020- | ···················· | ···················· | ····················· | ···················· | ···················· | ···················· |
| U | 84 | hCoV-19/USA/VA-DCLS-2046/2020\|EPI_ISL_677218\|2020- | ···················· | ···················· | ····················· | ···················· | ···················· | ···················· |
| U | 85 | hCoV-19/USA/VA-DCLS-2048/2020\|EPI_ISL_677219\|2020- | ···················· | ···················· | ····················· | ···················· | ···················· | ···················· |
| U | 86 | hCoV-19/USA/VA-DCLS-2049/2020\|EPI_ISL_677220\|2020- | ···················· | ···················· | ····················· | ···················· | ···················· | ···················· |
| U | 87 | hCoV-19/USA/VA-DCLS-2050/2020\|EPI_ISL_677221\|2020- | ···················· | ···················· | ····················· | ···················· | ···················· | ···················· |
| U | 88 | hCoV-19/USA/VA-DCLS-2051/2020\|EPI_ISL_677222\|2020- | ···················· | ···················· | ····················· | ···················· | ···················· | ···················· |
| U | 89 | hCoV-19/USA/VA-DCLS-2052/2020\|EPI_ISL_677223\|2020- | ···················· | ···················· | ····················· | ···················· | ···················· | ···················· |
| U | 90 | hCoV-19/USA/VA-DCLS-2054/2020\|EPI_ISL_677224\|2020- | ···················· | ···················· | ····················· | ···················· | ···················· | ···················· |
| U | 91 | hCoV-19/USA/VA-DCLS-2055/2020\|EPI_ISL_677225\|2020- | ···················· | ···················· | ····················· | ···················· | ···················· | ···················· |
| U | 92 | hCoV-19/USA/VA-DCLS-2056/2020\|EPI_ISL_677226\|2020- | ···················· | ···················· | ····················· | ···················· | ···················· | ···················· |
| U | 93 | hCoV-19/USA/VA-DCLS-2057/2020\|EPI_ISL_677227\|2020- | ···················· | ···················· | ····················· | ···················· | ···················· | ···················· |
| U | 94 | hCoV-19/France/ARA-15834/2020\|EPI_ISL_678500\|2020- | ···················· | ···················· | ····················· | ···················· | ···················· | ···················· |
| U | 95 | hCoV-19/France/ARA-16920/2020\|EPI_ISL_678501\|2020- | ···················· | ···················· | ····················· | ···················· | ···················· | ···················· |
| U | 96 | hCoV-19/France/ARA-18038/2020\|EPI_ISL_678502\|2020- | ···················· | ···················· | ····················· | ···················· | ···················· | ···················· |
| U | 97 | hCoV-19/France/ARA-18673/2020\|EPI_ISL_678503\|2020- | ···················· | ···················· | ····················· | ···················· | ···················· | ···················· |
| U | 98 | hCoV-19/France/ARA-193100/2020\|EPI_ISL_678504\|2020 | ···················· | ···················· | ····················· | ···················· | ···················· | ···················· |
| U | 99 | hCoV-19/France/ARA-19394/2020\|EPI_ISL_678505\|2020- | ···················· | ···················· | ····················· | ···················· | ···················· | ···················· |
| U | 100 | hCoV-19/France/ARA-2314/2020\|EPI_ISL_678506\|2020-0 | ···················· | ···················· | ····················· | ···················· | ···················· | ···················· |
| U | 101 | hCoV-19/France/ARA-2804/2020\|EPI_ISL_678507\|2020-0 | ···················· | ···················· | ····················· | ···················· | ···················· | ···················· |
| U | 102 | hCoV-19/France/ARA-37861/2020\|EPI_ISL_678508\|2020- | ···················· | ···················· | ····················· | ···················· | ···················· | ···················· |
| U | 103 | hCoV-19/USA/UT-UPHL-201207200/2020\|EPI_ISL_678516\| | ···················· | ···················· | ····················· | ···················· | ···················· | ···················· |
| U | 104 | hCoV-19/USA/UT-UPHL-201208274/2020\|EPI_ISL_678517\| | ···················· | ···················· | ····················· | ···················· | ···················· | ···················· |
| U | 105 | hCoV-19/USA/UT-UPHL-201212864/2020\|EPI_ISL_678519\| | ···················· | ···················· | ····················· | ···················· | ···················· | ···················· |
| U | 106 | hCoV-19/USA/UT-UPHL-201218231/2020\|EPI_ISL_678523\| | ···················· | ···················· | ····················· | ···················· | ···················· | ···················· |
| U | 107 | hCoV-19/USA/UT-UPHL-201222051/2020\|EPI_ISL_678525\| | ···················· | ···················· | ····················· | ···················· | ···················· | ···················· |
| U | 108 | hCoV-19/USA/UT-UPHL-201210677/2020\|EPI_ISL_678529\| | ···················· | ···················· | ····················· | ···················· | ···················· | ···················· |
| U | 109 | hCoV-19/USA/UT-UPHL-201232529/2020\|EPI_ISL_678530\| | ···················· | ···················· | ····················· | ···················· | ···················· | ···················· |
| U | 110 | hCoV-19/USA/UT-UPHL-201216248/2020\|EPI_ISL_678531\| | ···················· | ···················· | ····················· | ···················· | ···················· | ···················· |
| U | 111 | hCoV-19/USA/UT-UPHL-201225466/2020\|EPI_ISL_678532\| | ···················· | ···················· | ····················· | ···················· | ···················· | ···················· |
| U | 112 | hCoV-19/USA/UT-UPHL-201231390/2020\|EPI_ISL_678533\| | ···················· | ···················· | ····················· | ···················· | ···················· | ···················· |
| U | 113 | hCoV-19/France/ARA-27774/2020\|EPI_ISL_678534\|2020- | ···················· | ···················· | ····················· | ···················· | ···················· | ···················· |
| U | 114 | hCoV-19/France/ARA-29004/2020\|EPI_ISL_678535\|2020- | ···················· | ···················· | ····················· | ···················· | ···················· | ···················· |
| U | 115 | hCoV-19/France/ARA-30906/2020\|EPI_ISL_678536\|2020- | ···················· | ···················· | ····················· | ···················· | ···················· | ···················· |
| U | 116 | hCoV-19/France/ARA-31118/2020\|EPI_ISL_678537\|2020- | ···················· | ···················· | ····················· | ···················· | ···················· | ···················· |
| U | 117 | hCoV-19/France/ARA-31289/2020\|EPI_ISL_678538\|2020- | ···················· | ···················· | ····················· | ···················· | ···················· | ···················· |
| U | 118 | hCoV-19/France/ARA-32195/2020\|EPI_ISL_678539\|2020- | ···················· | ···················· | ····················· | ···················· | ···················· | ···················· |
| U | 119 | hCoV-19/France/ARA-32536/2020\|EPI_ISL_678540\|2020- | ···················· | ···················· | ····················· | ···················· | ···················· | ···················· |
| U | 120 | hCoV-19/France/ARA-32910/2020\|EPI_ISL_678541\|2020- | ···················· | ···················· | ····················· | ···················· | ···················· | ···················· |
| U | 121 | hCoV-19/France/ARA-33366/2020\|EPI_ISL_678542\|2020- | ···················· | ···················· | ····················· | ···················· | ···················· | ···················· |
| U | 122 | hCoV-19/France/ARA-36371/2020\|EPI_ISL_678543\|2020- | ···················· | ···················· | ····················· | ···················· | ···················· | ···················· |
| U | 123 | hCoV-19/France/ARA-36678/2020\|EPI_ISL_678544\|2020- | ···················· | ···················· | ····················· | ···················· | ···················· | ···················· |
| U | 124 | hCoV-19/France/ARA-38660/2020\|EPI_ISL_678545\|2020- | ···················· | ···················· | ····················· | ···················· | ···················· | ···················· |
| U | 125 | hCoV-19/France/ARA-40291/2020\|EPI_ISL_678546\|2020- | ···················· | ···················· | ····················· | ···················· | ···················· | ···················· |
| U | 126 | hCoV-19/France/ARA-40724/2020\|EPI_ISL_678547\|2020- | ···················· | ···················· | ····················· | ···················· | ···················· | ···················· |
| U | 127 | hCoV-19/England/MILK-B3BCA3/2020\|EPI_ISL_679802\|20 | ···················· | ···················· | ····················· | ···················· | ···················· | ···················· |
| U | 128 | hCoV-19/England/MILK-B3BCD0/2020\|EPI_ISL_679803\|20 | ···················· | ···················· | ····················· | ···················· | ···················· | ···················· |
| U | 129 | hCoV-19/England/MILK-B3BCEF/2020\|EPI_ISL_679804\|20 | ···················· | ···················· | ····················· | ···················· | ···················· | ···················· |
| U | 130 | hCoV-19/England/MILK-B3BCFE/2020\|EPI_ISL_679805\|20 | ···················· | ···················· | ····················· | ···················· | ···················· | ···················· |
| U | 131 | hCoV-19/England/MILK-B3BD28/2020\|EPI_ISL_679806\|20 | ···················· | ···················· | ····················· | ···················· | ···················· | ···················· |
| U | 132 | hCoV-19/England/MILK-B3BD64/2020\|EPI_ISL_679807\|20 | ···················· | ···················· | ····················· | ···················· | ······T············· | ···················· |
| U | 133 | hCoV-19/England/MILK-B3BDA0/2020\|EPI_ISL_679810\|20 | ···················· | ···················· | ····················· | ···················· | ···················· | ···················· |
| U | 134 | hCoV-19/England/MILK-B3BDDD/2020\|EPI_ISL_679811\|20 | ···················· | ···················· | ····················· | ···················· | ···················· | ···················· |
| U | 135 | hCoV-19/England/MILK-B3BDFB/2020\|EPI_ISL_679812\|20 | ···················· | ···················· | ····················· | ···················· | ···················· | ···················· |
| U | 136 | hCoV-19/England/MILK-B3C4E6/2020\|EPI_ISL_679813\|20 | ···················· | ···················· | ····················· | ···················· | ···················· | ···················· |
| U | 137 | hCoV-19/England/MILK-B3C501/2020\|EPI_ISL_679814\|20 | ···················· | ···················· | ····················· | ···················· | ···················· | ···················· |
| U | 138 | hCoV-19/England/MILK-B3C510/2020\|EPI_ISL_679815\|20 | ···················· | ···················· | ····················· | ···················· | ···················· | ···················· |
| U | 139 | hCoV-19/England/MILK-B3C589/2020\|EPI_ISL_679817\|20 | ···················· | ···················· | ····················· | ···················· | ······T············· | ···················· |
| U | 140 | hCoV-19/England/MILK-B3C5B6/2020\|EPI_ISL_679819\|20 | ···················· | ···················· | ····················· | ···················· | ···················· | ···················· |
| U | 141 | hCoV-19/England/MILK-B3C5E3/2020\|EPI_ISL_679820\|20 | ···················· | ···················· | ····················· | ···················· | ······T············· | ···················· |
| U | 142 | hCoV-19/England/MILK-B3CC0C/2020\|EPI_ISL_679821\|20 | ···················· | ···················· | ····················· | ···················· | ···················· | ···················· |
| U | 143 | hCoV-19/England/MILK-B3CC1B/2020\|EPI_ISL_679822\|20 | ···················· | ···················· | ····················· | ···················· | ···················· | ···················· |
| U | 144 | hCoV-19/England/MILK-B3CC39/2020\|EPI_ISL_679823\|20 | ···················· | ···················· | ····················· | ···················· | ···················· | ···················· |
| U | 145 | hCoV-19/England/MILK-B3CC57/2020\|EPI_ISL_679824\|20 | ···················· | ···················· | ····················· | ···················· | ···················· | ···················· |
| U | 146 | hCoV-19/England/MILK-B3CC75/2020\|EPI_ISL_679825\|20 | ···················· | ···················· | ····················· | ···················· | ···················· | ···················· |
| U | 147 | hCoV-19/England/MILK-B3CC84/2020\|EPI_ISL_679826\|20 | ···················· | ···················· | ····················· | ···················· | ···················· | ···················· |
| U | 148 | hCoV-19/England/MILK-B3CCB1/2020\|EPI_ISL_679828\|20 | ···················· | ···················· | ····················· | ···················· | ···················· | ···················· |
| U | 149 | hCoV-19/England/MILK-B3CCDF/2020\|EPI_ISL_679829\|20 | ···················· | ···················· | ····················· | ···················· | ···················· | ···················· |

| | | | | | | | |
|---|---|---|---|---|---|---|---|
| U | 150 hCoV-19/England/MILK-B3CCEE/2020\|EPI_ISL_679830\|20 | ···················· | ···················· | ····················· | ···················· | ···················· | ···················· |
| U | 151 hCoV-19/England/MILK-B3CD09/2020\|EPI_ISL_679831\|20 | ···················· | ···················· | ····················· | ···················· | ···················· | ···················· |
| U | 152 hCoV-19/England/MILK-B3D2DC/2020\|EPI_ISL_679832\|20 | ···················· | ···················· | ····················· | ···················· | ···················· | ···················· |
| U | 153 hCoV-19/England/MILK-B3D2EB/2020\|EPI_ISL_679833\|20 | ···················· | ···················· | ····················· | ···················· | ···················· | ···················· |
| U | 154 hCoV-19/England/MILK-B3D2FA/2020\|EPI_ISL_679834\|20 | ···················· | ···················· | ····················· | ···················· | ···················· | ···················· |
| U | 155 hCoV-19/England/MILK-B3D306/2020\|EPI_ISL_679835\|20 | ···················· | ···················· | ····················· | ···················· | ···················· | ···················· |
| U | 156 hCoV-19/England/MILK-B3D342/2020\|EPI_ISL_679836\|20 | ···················· | ···················· | ····················· | ···················· | ···················· | ···················· |
| U | 157 hCoV-19/England/MILK-B3D89E/2020\|EPI_ISL_679837\|20 | ···················· | ···················· | ····················· | ···················· | ···················· | ···················· |
| U | 158 hCoV-19/England/MILK-B3D8AD/2020\|EPI_ISL_679838\|20 | ···················· | ···················· | ····················· | ···················· | ···················· | ···················· |
| U | 159 hCoV-19/England/MILK-B3DFE4/2020\|EPI_ISL_679839\|20 | ···················· | ···················· | ····················· | ···················· | ······T············· | ···················· |
| U | 160 hCoV-19/England/MILK-B3DFF3/2020\|EPI_ISL_679840\|20 | ···················· | ···················· | ····················· | ···················· | ···················· | ···················· |
| U | 161 hCoV-19/England/MILK-B3E04A/2020\|EPI_ISL_679842\|20 | ···················· | ···················· | ····················· | ···················· | ···················· | ···················· |
| U | 162 hCoV-19/England/MILK-B3E059/2020\|EPI_ISL_679843\|20 | ···················· | ···················· | ····················· | ···················· | ···················· | ···················· |
| U | 163 hCoV-19/England/MILK-B3E077/2020\|EPI_ISL_679844\|20 | ···················· | ···················· | ····················· | ···················· | ···················· | ···················· |
| U | 164 hCoV-19/England/MILK-B3E095/2020\|EPI_ISL_679845\|20 | ···················· | ···················· | ····················· | ···················· | ······T············· | ···················· |
| U | 165 hCoV-19/England/MILK-B3E0A4/2020\|EPI_ISL_679846\|20 | ···················· | ···················· | ····················· | ···················· | ···················· | ···················· |
| U | 166 hCoV-19/England/MILK-B3E0B3/2020\|EPI_ISL_679847\|20 | ···················· | ···················· | ····················· | ···················· | ···················· | ···················· |
| U | 167 hCoV-19/England/MILK-B3E0E0/2020\|EPI_ISL_679848\|20 | ···················· | ···················· | ····················· | ···················· | ···················· | ···················· |
| U | 168 hCoV-19/England/MILK-B3E0FF/2020\|EPI_ISL_679849\|20 | ···················· | ···················· | ····················· | ···················· | ···················· | ···················· |
| U | 169 hCoV-19/England/MILK-B3E2CC/2020\|EPI_ISL_679850\|20 | ···················· | ···················· | ····················· | ···················· | ···················· | ···················· |
| U | 170 hCoV-19/England/MILK-B3E2DB/2020\|EPI_ISL_679851\|20 | ···················· | ···················· | ····················· | ···················· | ···················· | ···················· |
| U | 171 hCoV-19/England/MILK-B596FC/2020\|EPI_ISL_679858\|20 | ···················· | ···················· | ····················· | ···················· | ···················· | ···················· |
| U | 172 hCoV-19/England/MILK-B59832/2020\|EPI_ISL_679874\|20 | ···················· | ···················· | ····················· | ···················· | ···················· | ···················· |
| U | 173 hCoV-19/England/MILK-B5986F/2020\|EPI_ISL_679877\|20 | ···················· | ···················· | ····················· | ···················· | ···················· | ···················· |
| U | 174 hCoV-19/England/MILK-B5993F/2020\|EPI_ISL_679887\|20 | ···················· | ···················· | ····················· | ···················· | ···················· | ···················· |
| U | 175 hCoV-19/England/MILK-B59AF0/2020\|EPI_ISL_679907\|20 | ···················· | ···················· | ····················· | ···················· | ···················· | ···················· |
| U | 176 hCoV-19/England/MILK-B59B48/2020\|EPI_ISL_679910\|20 | ···················· | ···················· | ····················· | ···················· | ···················· | ···················· |
| U | 177 hCoV-19/England/MILK-B59B75/2020\|EPI_ISL_679912\|20 | ···················· | ···················· | ····················· | ···················· | ···················· | ···················· |
| U | 178 hCoV-19/England/MILK-B59C72/2020\|EPI_ISL_679922\|20 | ···················· | ···················· | ····················· | ···················· | ···················· | ···················· |
| U | 179 hCoV-19/Wales/PHWC-48E07F/2020\|EPI_ISL_680800\|2020 | ···················· | ···················· | ····················· | ···················· | ···················· | ···················· |
| U | 180 hCoV-19/Wales/PHWC-48E08E/2020\|EPI_ISL_680801\|2020 | ···················· | ···················· | ····················· | ···················· | ···················· | ···················· |
| U | 181 hCoV-19/Wales/PHWC-48E0AC/2020\|EPI_ISL_680803\|2020 | ···················· | ···················· | ····················· | ···················· | ···················· | ···················· |
| U | 182 hCoV-19/Wales/PHWC-48E0BB/2020\|EPI_ISL_680804\|2020 | ···················· | ···················· | ····················· | ···················· | ···················· | ···················· |
| U | 183 hCoV-19/Wales/PHWC-48E0CA/2020\|EPI_ISL_680805\|2020 | ···················· | ···················· | ····················· | ···················· | ···················· | ···················· |
| U | 184 hCoV-19/Wales/PHWC-48E0D9/2020\|EPI_ISL_680806\|2020 | ···················· | ···················· | ····················· | ···················· | ···················· | ···················· |
| U | 185 hCoV-19/Wales/PHWC-48E0E8/2020\|EPI_ISL_680807\|2020 | ···················· | ···················· | ····················· | ···················· | ···················· | ···················· |
| U | 186 hCoV-19/Wales/PHWC-48E103/2020\|EPI_ISL_680809\|2020 | ···················· | ···················· | ····················· | ···················· | ···················· | ···················· |
| U | 187 hCoV-19/Wales/PHWC-48E112/2020\|EPI_ISL_680810\|2020 | ···················· | ···················· | ····················· | ···················· | ···················· | ···················· |
| U | 188 hCoV-19/Wales/PHWC-48E121/2020\|EPI_ISL_680811\|2020 | ···················· | ···················· | ····················· | ···················· | ···················· | ···················· |
| U | 189 hCoV-19/Wales/PHWC-48E130/2020\|EPI_ISL_680812\|2020 | ···················· | ···················· | ····················· | ···················· | ···················· | ···················· |
| U | 190 hCoV-19/Wales/PHWC-48E14F/2020\|EPI_ISL_680813\|2020 | ···················· | ···················· | ····················· | ···················· | ···················· | ···················· |
| U | 191 hCoV-19/Wales/PHWC-48E16D/2020\|EPI_ISL_680815\|2020 | ···················· | ···················· | ····················· | ···················· | ···················· | ···················· |
| U | 192 hCoV-19/Wales/PHWC-48E17C/2020\|EPI_ISL_680816\|2020 | ···················· | ···················· | ····················· | ···················· | ···················· | ···················· |
| U | 193 hCoV-19/Wales/PHWC-48E18B/2020\|EPI_ISL_680817\|2020 | ···················· | ···················· | ····················· | ···················· | ···················· | ···················· |
| U | 194 hCoV-19/Wales/PHWC-48E19A/2020\|EPI_ISL_680818\|2020 | ···················· | ···················· | ····················· | ···················· | ···················· | ···················· |
| U | 195 hCoV-19/Wales/PHWC-48E1B8/2020\|EPI_ISL_680819\|2020 | ···················· | ···················· | ····················· | ···················· | ···················· | ···················· |
| U | 198 hCoV-19/Wales/PHWC-48E1F4/2020\|EPI_ISL_680823\|2020 | ···················· | ···················· | ····················· | ···················· | ···················· | ···················· |
| U | 199 hCoV-19/Wales/PHWC-48E200/2020\|EPI_ISL_680824\|2020 | ···················· | ···················· | ····················· | ···················· | ···················· | ···················· |
| U | 200 hCoV-19/Wales/PHWC-48E21F/2020\|EPI_ISL_680825\|2020 | ···················· | ···················· | ····················· | ···················· | ···················· | ···················· |
| U | 201 hCoV-19/Wales/PHWC-48E23D/2020\|EPI_ISL_680827\|2020 | ···················· | ···················· | ····················· | ···················· | ···················· | ···················· |
| U | 202 hCoV-19/Wales/PHWC-48E25B/2020\|EPI_ISL_680829\|2020 | ···················· | ······A············· | ····················· | ···················· | ···················· | ···················· |
| U | 203 hCoV-19/Wales/PHWC-48E26A/2020\|EPI_ISL_680830\|2020 | ···················· | ···················· | ····················· | ···················· | ···················· | ···················· |
| U | 206 hCoV-19/Wales/PHWC-48E2A6/2020\|EPI_ISL_680834\|2020 | ···················· | ···················· | ····················· | ···················· | ···A················ | ···················· |
| U | 208 hCoV-19/Wales/PHWC-48E2C4/2020\|EPI_ISL_680836\|2020 | ···················· | ···················· | ····················· | ···················· | ···················· | ···················· |
| U | 209 hCoV-19/Wales/PHWC-48E2E2/2020\|EPI_ISL_680838\|2020 | ···················· | ···················· | ····················· | ···················· | ···················· | ···················· |
| U | 210 hCoV-19/Wales/PHWC-48E2F1/2020\|EPI_ISL_680839\|2020 | ···················· | ···················· | ····················· | ···················· | ···················· | ···················· |
| U | 211 hCoV-19/Wales/PHWC-48E30D/2020\|EPI_ISL_680840\|2020 | ···················· | ···················· | ····················· | ···················· | ···················· | ···················· |
| U | 212 hCoV-19/Wales/PHWC-48E31C/2020\|EPI_ISL_680841\|2020 | ···················· | ······A············· | ····················· | ···················· | ···················· | ···················· |
| U | 213 hCoV-19/Wales/PHWC-48E32B/2020\|EPI_ISL_680842\|2020 | ···················· | ······A············· | ····················· | ···················· | ···················· | ···················· |
| U | 214 hCoV-19/Wales/PHWC-48E33A/2020\|EPI_ISL_680843\|2020 | ···················· | ···················· | ····················· | ···················· | ···················· | ···················· |
| U | 215 hCoV-19/Wales/PHWC-48E349/2020\|EPI_ISL_680844\|2020 | ···················· | ···················· | ····················· | ···················· | ···················· | ···················· |
| U | 216 hCoV-19/Wales/PHWC-48E358/2020\|EPI_ISL_680845\|2020 | ···················· | ···················· | ····················· | ···················· | ···A················ | ···················· |
| U | 218 hCoV-19/Wales/PHWC-48E394/2020\|EPI_ISL_680849\|2020 | ···················· | ···················· | ····················· | ···················· | ···················· | ···················· |
| U | 219 hCoV-19/Wales/PHWC-48E3C1/2020\|EPI_ISL_680851\|2020 | ···················· | ···················· | ····················· | ···················· | ···················· | ···················· |
| U | 220 hCoV-19/Wales/PHWC-48E3D0/2020\|EPI_ISL_680852\|2020 | ···················· | ···················· | ····················· | ···················· | ···················· | ···················· |
| U | 221 hCoV-19/Wales/PHWC-48E3EF/2020\|EPI_ISL_680853\|2020 | ···················· | ···················· | ····················· | ···················· | ···················· | ···················· |
| U | 222 hCoV-19/Wales/PHWC-48E3FE/2020\|EPI_ISL_680854\|2020 | ···················· | ···················· | ····················· | ···················· | ···················· | ···················· |
| U | 223 hCoV-19/Wales/PHWC-48E40A/2020\|EPI_ISL_680855\|2020 | ···················· | ···················· | ····················· | ···················· | ···················· | ···················· |
| U | 226 hCoV-19/Wales/PHWC-48E455/2020\|EPI_ISL_680860\|2020 | ···················· | ···················· | ····················· | ···················· | ···················· | ···················· |
| U | 227 hCoV-19/Wales/PHWC-48E464/2020\|EPI_ISL_680861\|2020 | ···················· | ···················· | ····················· | ···················· | ···················· | ···················· |
| U | 228 hCoV-19/Wales/PHWC-48E473/2020\|EPI_ISL_680862\|2020 | ···················· | ···················· | ····················· | ···················· | ···················· | ···················· |
| U | 229 hCoV-19/Wales/PHWC-48E482/2020\|EPI_ISL_680863\|2020 | ···················· | ···················· | ····················· | ···················· | ···················· | ···················· |
| U | 231 hCoV-19/Wales/PHWC-48E4CE/2020\|EPI_ISL_680866\|2020 | ···················· | ···················· | ····················· | ···················· | ···················· | ···················· |
| U | 234 hCoV-19/Wales/PHWC-48E4FB/2020\|EPI_ISL_680869\|2020 | ···················· | ···················· | ····················· | ···················· | ···················· | ···················· |
| U | 235 hCoV-19/Wales/PHWC-48E701/2020\|EPI_ISL_680900\|2020 | ···················· | ···················· | ····················· | ···················· | ···················· | ···················· |
| U | 236 hCoV-19/Wales/PHWC-48E710/2020\|EPI_ISL_680901\|2020 | ···················· | ···················· | ····················· | ···················· | ···················· | ···················· |
| U | 237 hCoV-19/Wales/PHWC-48E72F/2020\|EPI_ISL_680902\|2020 | ···················· | ···················· | ····················· | ···················· | ···················· | ···················· |
| U | 238 hCoV-19/Wales/PHWC-48E73E/2020\|EPI_ISL_680903\|2020 | ···················· | ···················· | ····················· | ···················· | ···················· | N··················· |
| U | 239 hCoV-19/Wales/PHWC-48E74D/2020\|EPI_ISL_680904\|2020 | ···················· | ···················· | ····················· | ···················· | ···················· | ···················· |
| U | 240 hCoV-19/Wales/PHWC-48E75C/2020\|EPI_ISL_680905\|2020 | ···················· | ···················· | ····················· | ···················· | ···················· | ···················· |

U 242 hCoV-19/Wales/PHWC-48E77A/2020|EPI_ISL_680907|2020 ···················· ···················· ····················· ···················· ···················· ····················
U 243 hCoV-19/Wales/PHWC-48E789/2020|EPI_ISL_680908|2020 ···················· ···················· ····················· ···················· ···················· ····················
U 244 hCoV-19/Wales/PHWC-48E798/2020|EPI_ISL_680909|2020 ···················· ···················· ····················· ···················· ···················· ····················
U 245 hCoV-19/mink/Denmark/mDK-235/2020|EPI_ISL_683085|2 ···················· ·A·················· ····················· ···················· ···················· ····················
U 246 hCoV-19/mink/Denmark/mDK-238/2020|EPI_ISL_683087|2 ···················· ·A·················· ····················· ···················· ···················· ····················
U 247 hCoV-19/mink/Denmark/mDK-241/2020|EPI_ISL_683090|2 ···················· ·N·················· ····················· ···················· ···················· ····················
U 248 hCoV-19/mink/Denmark/mDK-266/2020|EPI_ISL_683119|2 ···················· ·A·················· ····················· ···················· ···················· ····················
U 249 hCoV-19/mink/Denmark/mDK-273/2020|EPI_ISL_683125|2 ···················· ·N·················· ····················· ···················· ···················· ····················
U 250 hCoV-19/mink/Denmark/mDK-288/2020|EPI_ISL_683138|2 ···················· ·A·················· ····················· ···················· ···················· ····················
U 251 hCoV-19/mink/Denmark/mDK-323/2020|EPI_ISL_683172|2 ···················· ·A·················· ····················· ···················· ···················· ····················
U 252 hCoV-19/mink/Denmark/mDK-326/2020|EPI_ISL_683175|2 ···················· ····A··············· ····················· ···················· ···················· ····················
U 253 hCoV-19/mink/Denmark/mDK-327/2020|EPI_ISL_683176|2 ···················· ····A··············· ····················· ···················· ···················· ····················
U 254 hCoV-19/mink/Denmark/mDK-328/2020|EPI_ISL_683177|2 ···················· ····A··············· ····················· ···················· ···················· ····················
U 255 hCoV-19/mink/Denmark/mDK-430/2020|EPI_ISL_683188|2 ···················· ·A·················· ····················· ···················· ···················· ····················
U 256 hCoV-19/mink/Denmark/mDK-432/2020|EPI_ISL_683189|2 ···················· ·A·················· ····················· ···················· ···················· ····················
U 257 hCoV-19/England/MILK-B957F1/2020|EPI_ISL_673601|20 ···················· ···················· ····················· ···················· ···················· ····················
U 258 hCoV-19/England/CAMC-B9946F/2020|EPI_ISL_673602|20 ···················· ···················· ····················· ···················· ···················· ····················
U 259 hCoV-19/England/MILK-B9532A/2020|EPI_ISL_673603|20 ···················· ·A·················· ····················· ···················· ···················· ···A················
U 260 hCoV-19/England/CAMC-B9873A/2020|EPI_ISL_673604|20 ···················· ···················· ····················· ···················· ···················· ····················
U 261 hCoV-19/England/CAMC-B9A194/2020|EPI_ISL_673605|20 ···················· ···················· ····················· ···················· ···················· ····················
U 262 hCoV-19/England/CAMC-B9914A/2020|EPI_ISL_673606|20 ···················· ···················· ····················· ···················· ···················· ····················
U 263 hCoV-19/England/CAMC-B98952/2020|EPI_ISL_673609|20 ···················· ···················· ····················· ···················· ···················· ····················
U 264 hCoV-19/England/CAMC-B98DB0/2020|EPI_ISL_673610|20 ···················· ···················· ····················· ···················· ···················· ····················
U 265 hCoV-19/England/CAMC-B98336/2020|EPI_ISL_673611|20 ···················· ···················· ····················· ···················· ···················· ····················
U 266 hCoV-19/England/CAMC-B9AE7F/2020|EPI_ISL_673613|20 ···················· ···················· ····················· ···················· ···················· ····················
U 267 hCoV-19/England/CAMC-B982DF/2020|EPI_ISL_673614|20 ···················· ···················· ····················· ···················· ···················· ····················
U 268 hCoV-19/England/CAMC-B9A352/2020|EPI_ISL_673615|20 ···················· ···················· ····················· ···················· ···················· ····················
U 269 hCoV-19/England/CAMC-B98DA1/2020|EPI_ISL_673617|20 ···················· ···················· ····················· ···················· ···················· ····················
U 270 hCoV-19/England/CAMC-B983AF/2020|EPI_ISL_673619|20 ···················· ···················· ····················· ···················· ···················· ····················
U 271 hCoV-19/England/CAMC-B9A1C1/2020|EPI_ISL_673620|20 ···················· ···················· ····················· ···················· ···················· ····················
U 272 hCoV-19/England/MILK-B94BE7/2020|EPI_ISL_673621|20 ···················· ···················· ····················· ···················· ···················· ····················
U 273 hCoV-19/England/CAMC-B98DFC/2020|EPI_ISL_673624|20 ···················· ···················· ····················· ···················· ···················· ····················
U 274 hCoV-19/England/CAMC-B98819/2020|EPI_ISL_673625|20 ···················· ···················· ····················· ···················· ···················· ····················
U 275 hCoV-19/England/CAMC-B98CE0/2020|EPI_ISL_673626|20 ···················· ···················· ····················· ···················· ···················· ····················
U 276 hCoV-19/England/CAMC-B9AECA/2020|EPI_ISL_673628|20 ···················· ···················· ····················· ···················· ···················· ····················
U 277 hCoV-19/England/CAMC-B98C0E/2020|EPI_ISL_673630|20 ···················· ···················· ····················· ···················· ···················· ····················
U 278 hCoV-19/England/CAMC-B994BA/2020|EPI_ISL_673632|20 ···················· ·A·················· ····················· ···················· ···················· ···A················
U 279 hCoV-19/England/MILK-B952B4/2020|EPI_ISL_673633|20 ···················· ···················· ····················· ···················· ···················· ····················
U 280 hCoV-19/England/CAMC-B99924/2020|EPI_ISL_673637|20 ···················· ···················· ····················· ···················· ···················· ····················
U 281 hCoV-19/England/CAMC-B989AD/2020|EPI_ISL_673640|20 ···················· ···················· ····················· ···················· ···················· ····················
U 282 hCoV-19/England/MILK-B950E7/2020|EPI_ISL_673642|20 ···················· ···················· ····················· ···················· ···················· ····················
U 283 hCoV-19/England/CAMC-B9AE60/2020|EPI_ISL_673643|20 ···················· ···················· ····················· ···················· ···················· ····················
U 284 hCoV-19/England/CAMC-B985A9/2020|EPI_ISL_673644|20 ···················· ···················· ····················· ···················· ···················· ····················
U 285 hCoV-19/England/MILK-B948FF/2020|EPI_ISL_673646|20 ···················· ···················· ····················· ···················· ···················· ····················
U 286 hCoV-19/England/CAMC-B98D74/2020|EPI_ISL_673647|20 ···················· ···················· ····················· ···················· ···················· ····················
U 287 hCoV-19/England/CAMC-B98E71/2020|EPI_ISL_673648|20 ···················· ···················· ····················· ···················· ···················· ····················
U 288 hCoV-19/England/CAMC-B99F04/2020|EPI_ISL_673649|20 ···················· ···················· ····················· ···················· ···················· ····················
U 289 hCoV-19/England/CAMC-B988CE/2020|EPI_ISL_673651|20 ···················· ···················· ····················· ···················· ···················· ····················
U 290 hCoV-19/England/CAMC-B98D29/2020|EPI_ISL_673653|20 ···················· ·A·················· ····················· ···················· ···················· ···A················
U 291 hCoV-19/England/CAMC-B98BF2/2020|EPI_ISL_673654|20 ···················· ·A·················· ····················· ···················· ···················· ···A················
U 292 hCoV-19/England/MILK-B9508D/2020|EPI_ISL_673655|20 ···················· ···················· ····················· ···················· ···················· ····················
U 293 hCoV-19/England/CAMC-B9AEBB/2020|EPI_ISL_673656|20 ···················· ···················· ····················· ···················· ···················· ····················
U 294 hCoV-19/England/MILK-B95287/2020|EPI_ISL_673657|20 ···················· ···················· ····················· ···················· ···················· ····················
U 295 hCoV-19/England/CAMC-B983CD/2020|EPI_ISL_673658|20 ···················· ·A·················· ····················· ···················· ···················· ···A················
U 296 hCoV-19/England/MILK-B951F3/2020|EPI_ISL_673659|20 ···················· ···················· ····················· ···················· ···················· ····················
U 297 hCoV-19/England/CAMC-B9A6A4/2020|EPI_ISL_673660|20 ···················· ···················· ····················· ···················· ···················· ····················
U 298 hCoV-19/England/MILK-B950AB/2020|EPI_ISL_673663|20 ···················· ···················· ····················· ···················· ···················· ····················
U 299 hCoV-19/England/CAMC-B984F7/2020|EPI_ISL_673664|20 ···················· ···················· ····················· ···················· ···················· ····················
U 300 hCoV-19/England/CAMC-B9A62C/2020|EPI_ISL_673665|20 ···················· ···················· ····················· ···················· ···················· ····················
U 301 hCoV-19/England/CAMC-B988A0/2020|EPI_ISL_673666|20 ···················· ···················· ····················· ···················· ···················· ····················
U 302 hCoV-19/England/CAMC-B9A316/2020|EPI_ISL_673667|20 ···················· ···················· ····················· ···················· ···················· ····················
U 303 hCoV-19/England/CAMC-B9A6E0/2020|EPI_ISL_673668|20 ···················· ···················· ····················· ···················· ···················· ····················
U 304 hCoV-19/England/CAMC-B9855E/2020|EPI_ISL_673669|20 ···················· ···················· ····················· ···················· ···················· ····················
U 305 hCoV-19/England/CAMC-B988DD/2020|EPI_ISL_673670|20 ···················· ···················· ····················· ···················· ···················· ····················
U 306 hCoV-19/England/CAMC-B99E9E/2020|EPI_ISL_673671|20 ···················· ···················· ····················· ···················· ···················· ····················
U 307 hCoV-19/England/CAMC-B98DDE/2020|EPI_ISL_673672|20 ···················· ···················· ····················· ···················· ···················· ····················
U 308 hCoV-19/England/MILK-B95357/2020|EPI_ISL_673673|20 ···················· ···················· ····················· ···················· ···················· ····················
U 309 hCoV-19/England/CAMC-B98309/2020|EPI_ISL_673674|20 ···················· ···················· ····················· ···················· ······T············· ····················
U 310 hCoV-19/England/CAMC-B99177/2020|EPI_ISL_673675|20 ···················· ···················· ····················· ···················· ···················· ····················
U 311 hCoV-19/England/CAMC-B999AC/2020|EPI_ISL_673678|20 ···················· ···················· ····················· ···················· ···················· ····················
U 312 hCoV-19/England/CAMC-B9A61D/2020|EPI_ISL_673679|20 ···················· ···················· ····················· ···················· ···················· ····················
U 313 hCoV-19/England/CAMC-B999CA/2020|EPI_ISL_673680|20 ···················· ···················· ····················· ···················· ···················· ····················
U 314 hCoV-19/England/CAMC-B98A6E/2020|EPI_ISL_673682|20 ···················· ···················· ····················· ···················· ···················· ····················
U 315 hCoV-19/England/CAMC-B9905C/2020|EPI_ISL_673683|20 ···················· ···················· ····················· ···················· ···················· ····················
U 316 hCoV-19/England/CAMC-B98D47/2020|EPI_ISL_673684|20 ···················· ···················· ····················· ···················· ···················· ····················
U 317 hCoV-19/England/CAMC-B9870D/2020|EPI_ISL_673685|20 ···················· ···················· ····················· ···················· ···················· ····················
U 318 hCoV-19/England/CAMC-B98521/2020|EPI_ISL_673687|20 ···················· ···················· ····················· ···················· ···················· ····················
U 319 hCoV-19/England/MILK-B94947/2020|EPI_ISL_673688|20 ···················· ···················· ····················· ···················· ···················· ····················
U 320 hCoV-19/England/CAMC-B9871C/2020|EPI_ISL_673689|20 ···················· ···················· ····················· ···················· ···················· ····················
U 321 hCoV-19/England/CAMC-B98D65/2020|EPI_ISL_673690|20 ···················· ·A·················· ····················· ···················· ···················· ···A················

| | | | | | | | | |
|---|---|---|---|---|---|---|---|---|
| U | 322 | hCoV-19/England/CAMC-B99F40/2020\|EPI_ISL_673693\|20 | ···················· | ···················· | ····················· | ···················· | ···················· | ···················· |
| U | 323 | hCoV-19/England/CAMC-B9854F/2020\|EPI_ISL_673694\|20 | ···················· | ···················· | ····················· | ···················· | ···················· | ···················· |
| U | 324 | hCoV-19/England/CAMC-B99906/2020\|EPI_ISL_673695\|20 | ···················· | ···················· | ····················· | ···················· | ···················· | ···················· |
| U | 325 | hCoV-19/England/MILK-B9531B/2020\|EPI_ISL_673696\|20 | ···················· | ···················· | ····················· | ···················· | ···················· | ···················· |
| U | 326 | hCoV-19/England/CAMC-B9AF7C/2020\|EPI_ISL_673697\|20 | ···················· | ···················· | ····················· | ···················· | ···················· | ···················· |
| U | 327 | hCoV-19/England/CAMC-B98A5F/2020\|EPI_ISL_673698\|20 | ···················· | ···················· | ····················· | ···················· | ···················· | ···················· |
| U | 328 | hCoV-19/England/CAMC-B99915/2020\|EPI_ISL_673699\|20 | ···················· | ···················· | ····················· | ···················· | ···················· | ···················· |
| U | 329 | hCoV-19/England/CAMC-980B7/2020\|EPI_ISL_674910\|20 | ···················· | ···················· | ····················· | ···················· | ···················· | ···················· |
| U | 330 | hCoV-19/England/MILK-B959BF/2020\|EPI_ISL_674911\|20 | ···················· | ···················· | ····················· | ···················· | ···················· | ···················· |
| U | 331 | hCoV-19/England/MILK-B972A3/2020\|EPI_ISL_674914\|20 | ···················· | ···················· | ····················· | ···················· | ···················· | ···················· |
| U | 332 | hCoV-19/England/MILK-B9713D/2020\|EPI_ISL_674915\|20 | ···················· | ···················· | ····················· | ···················· | ···················· | ···················· |
| U | 333 | hCoV-19/England/MILK-B95ABC/2020\|EPI_ISL_674916\|20 | ···················· | ···················· | ····················· | ···················· | ···················· | ···················· |
| U | 334 | hCoV-19/England/CAMC-B98187/2020\|EPI_ISL_674917\|20 | ···················· | ···················· | ····················· | ···················· | ···················· | ···················· |
| U | 335 | hCoV-19/England/MILK-B97513/2020\|EPI_ISL_674918\|20 | ···················· | ···················· | ····················· | ···················· | ···················· | ···················· |
| U | 336 | hCoV-19/England/CAMC-B984CA/2020\|EPI_ISL_674919\|20 | ···················· | ···················· | ····················· | ···················· | ······T············· | ···················· |
| U | 337 | hCoV-19/England/MILK-B95E56/2020\|EPI_ISL_674921\|20 | ···················· | ·A·················· | ····················· | ···················· | ···················· | ···A················ |
| U | 338 | hCoV-19/England/CAMC-B97F8E/2020\|EPI_ISL_674922\|20 | ···················· | ···················· | ····················· | ···················· | ···················· | ···················· |
| U | 339 | hCoV-19/England/CAMC-B9807B/2020\|EPI_ISL_674923\|20 | ···················· | ···················· | ····················· | ···················· | ···················· | ···················· |
| U | 340 | hCoV-19/England/CAMC-B9806C/2020\|EPI_ISL_674924\|20 | ···················· | ···················· | ····················· | ···················· | ···················· | ···················· |
| U | 341 | hCoV-19/England/CAMC-B9811E/2020\|EPI_ISL_674926\|20 | ···················· | ·A·················· | ····················· | ···················· | ···················· | ···A················ |
| U | 342 | hCoV-19/England/MILK-B97373/2020\|EPI_ISL_674928\|20 | ···················· | ···················· | ····················· | ···················· | ···················· | ···················· |
| U | 343 | hCoV-19/England/MILK-B95E38/2020\|EPI_ISL_674929\|20 | ···················· | ···················· | ····················· | ···················· | ···················· | ···················· |
| U | 344 | hCoV-19/England/MILK-B95E0B/2020\|EPI_ISL_674930\|20 | ···················· | ···················· | ····················· | ···················· | ···················· | ···················· |
| U | 345 | hCoV-19/England/MILK-B97276/2020\|EPI_ISL_674931\|20 | ···················· | ···················· | ····················· | ···················· | ···················· | ···················· |
| U | 346 | hCoV-19/England/MILK-B97258/2020\|EPI_ISL_674932\|20 | ···················· | ···················· | ····················· | ···················· | ···················· | ···················· |
| U | 347 | hCoV-19/England/CAMC-B98688/2020\|EPI_ISL_674933\|20 | ···················· | ···················· | ····················· | ···················· | ···················· | ···················· |
| U | 348 | hCoV-19/England/MILK-B95A25/2020\|EPI_ISL_674934\|20 | ···················· | ···················· | ····················· | ···················· | ···················· | ···················· |
| U | 349 | hCoV-19/England/MILK-B95E74/2020\|EPI_ISL_674935\|20 | ···················· | ···················· | ····················· | ···················· | ···················· | ···················· |
| U | 350 | hCoV-19/England/MILK-B96E28/2020\|EPI_ISL_674937\|20 | ···················· | ···················· | ····················· | ···················· | ···················· | ···················· |
| U | 351 | hCoV-19/England/MILK-B9621D/2020\|EPI_ISL_674938\|20 | ···················· | ···················· | ····················· | ···················· | ···················· | ···················· |
| U | 352 | hCoV-19/England/CAMC-B984D9/2020\|EPI_ISL_674940\|20 | ···················· | ···················· | ····················· | ···················· | ···················· | ···················· |
| U | 353 | hCoV-19/England/MILK-B9656F/2020\|EPI_ISL_674943\|20 | ···················· | ···················· | ····················· | ···················· | ······T············· | ···················· |
| U | 354 | hCoV-19/England/CAMC-B97F7F/2020\|EPI_ISL_674944\|20 | ···················· | ···················· | ····················· | ···················· | ···················· | ···················· |
| U | 355 | hCoV-19/England/MILK-B95982/2020\|EPI_ISL_674945\|20 | ···················· | ···················· | ····················· | ···················· | ···················· | ···A················ |
| U | 356 | hCoV-19/England/CAMC-B97FBB/2020\|EPI_ISL_674946\|20 | ···················· | ···················· | ····················· | ···················· | ···················· | ···················· |
| U | 357 | hCoV-19/England/MILK-B964BD/2020\|EPI_ISL_674947\|20 | ···················· | ···················· | ····················· | ···················· | ···················· | ···················· |
| U | 358 | hCoV-19/England/MILK-B9706D/2020\|EPI_ISL_674948\|20 | ···················· | ···················· | ····················· | ···················· | ···················· | ···················· |
| U | 359 | hCoV-19/England/CAMC-B98266/2020\|EPI_ISL_674950\|20 | ···················· | ···················· | ····················· | ···················· | ···················· | ···················· |
| U | 360 | hCoV-19/England/CAMC-B986C4/2020\|EPI_ISL_674951\|20 | ···················· | ·A·················· | ····················· | ···················· | ···················· | ···A················ |
| U | 361 | hCoV-19/England/CAMC-B984BB/2020\|EPI_ISL_674952\|20 | ···················· | ···················· | ····················· | ···················· | ···················· | ···················· |
| U | 362 | hCoV-19/England/CAMC-B97FE8/2020\|EPI_ISL_674953\|20 | ···················· | ···················· | ····················· | ···················· | ···················· | ···················· |
| U | 363 | hCoV-19/England/MILK-B95CB6/2020\|EPI_ISL_674954\|20 | ···················· | ···················· | ····················· | ···················· | ···················· | ···················· |
| U | 364 | hCoV-19/England/MILK-B977D1/2020\|EPI_ISL_674956\|20 | ···················· | ···················· | ····················· | ···················· | ···················· | ···················· |
| U | 365 | hCoV-19/England/CAMC-B981C3/2020\|EPI_ISL_674957\|20 | ···················· | ···················· | ····················· | ···················· | ···················· | ···················· |
| U | 366 | hCoV-19/England/MILK-B9780B/2020\|EPI_ISL_674958\|20 | ···················· | ···················· | ····················· | ···················· | ···················· | ···················· |
| U | 367 | hCoV-19/England/CAMC-B98293/2020\|EPI_ISL_674959\|20 | ···················· | ···················· | ····················· | ···················· | ···················· | ···················· |
| U | 368 | hCoV-19/England/MILK-B96FDA/2020\|EPI_ISL_674960\|20 | ···················· | ···················· | ····················· | ···················· | ···················· | ···················· |
| U | 369 | hCoV-19/England/MILK-B974DA/2020\|EPI_ISL_674961\|20 | ···················· | ···················· | ····················· | ···················· | ···················· | ···················· |
| U | 370 | hCoV-19/England/CAMC-B980F3/2020\|EPI_ISL_674962\|20 | ···················· | ···················· | ····················· | ···················· | ···················· | ···················· |
| U | 371 | hCoV-19/England/MILK-B959EC/2020\|EPI_ISL_674963\|20 | ···················· | ···················· | ····················· | ···················· | ···················· | ···················· |
| U | 372 | hCoV-19/England/MILK-B9723A/2020\|EPI_ISL_674964\|20 | ···················· | ···················· | ····················· | ···················· | ···················· | ···················· |
| U | 373 | hCoV-19/England/MILK-B97021/2020\|EPI_ISL_674965\|20 | ···················· | ···················· | ····················· | ···················· | ···················· | ···················· |
| U | 374 | hCoV-19/England/MILK-B96DC1/2020\|EPI_ISL_674966\|20 | ···················· | ···················· | ····················· | ···················· | ···················· | ···················· |
| U | 375 | hCoV-19/England/MILK-B9704F/2020\|EPI_ISL_674967\|20 | ···················· | ·A·················· | ····················· | ···················· | ···················· | ···················· |
| U | 376 | hCoV-19/England/MILK-B96BE5/2020\|EPI_ISL_674968\|20 | ···················· | ···················· | ····················· | ···················· | ···················· | ···················· |
| U | 377 | hCoV-19/England/MILK-B97795/2020\|EPI_ISL_674969\|20 | ···················· | ···················· | ····················· | ···················· | ······T············· | ···················· |
| U | 378 | hCoV-19/England/MILK-B9765C/2020\|EPI_ISL_674970\|20 | ···················· | ···················· | ····················· | ···················· | ···················· | ···················· |
| U | 379 | hCoV-19/England/MILK-B96C2E/2020\|EPI_ISL_674971\|20 | ···················· | ···················· | ····················· | ···················· | ···················· | ···················· |
| U | 380 | hCoV-19/England/CAMC-B986A6/2020\|EPI_ISL_674972\|20 | ···················· | ···················· | ····················· | ···················· | ···················· | ···················· |
| U | 381 | hCoV-19/England/CAMC-B980D5/2020\|EPI_ISL_674973\|20 | ···················· | ···················· | ····················· | ···················· | ···················· | ···················· |
| U | 382 | hCoV-19/England/MILK-B96259/2020\|EPI_ISL_674974\|20 | ···················· | ···················· | ····················· | ···················· | ···················· | ···················· |
| U | 383 | hCoV-19/England/MILK-B95928/2020\|EPI_ISL_674975\|20 | ···················· | ···················· | ····················· | ···················· | ···················· | ···················· |
| U | 384 | hCoV-19/England/MILK-B95964/2020\|EPI_ISL_674976\|20 | ···················· | ···················· | ····················· | ···················· | ···················· | ···A················ |
| U | 385 | hCoV-19/England/CAMC-B98099/2020\|EPI_ISL_674977\|20 | ···················· | ···················· | ····················· | ···················· | ···················· | ···················· |
| U | 386 | hCoV-19/England/MILK-B95ADA/2020\|EPI_ISL_674978\|20 | ···················· | ···················· | ····················· | ···················· | ···················· | ···················· |
| U | 387 | hCoV-19/England/MILK-B961E3/2020\|EPI_ISL_674979\|20 | ···················· | ···················· | ····················· | ···················· | ···················· | ···················· |
| U | 388 | hCoV-19/England/MILK-B96C1F/2020\|EPI_ISL_674980\|20 | ···················· | ···················· | ····················· | ···················· | ···················· | ···················· |
| U | 389 | hCoV-19/England/MILK-B971F1/2020\|EPI_ISL_674981\|20 | ···················· | ···················· | ····················· | ···················· | ···················· | ···················· |
| U | 390 | hCoV-19/England/MILK-B97364/2020\|EPI_ISL_674982\|20 | ···················· | ···················· | ····················· | ···················· | ···················· | ···················· |
| U | 391 | hCoV-19/England/CAMC-B98442/2020\|EPI_ISL_674983\|20 | ···················· | ···················· | ····················· | ···················· | ···················· | ···················· |
| U | 392 | hCoV-19/England/MILK-B96C00/2020\|EPI_ISL_674984\|20 | ···················· | ···················· | ····················· | ···················· | ···················· | ···················· |
| U | 393 | hCoV-19/England/MILK-B96D94/2020\|EPI_ISL_674985\|20 | ···················· | ···················· | ····················· | ···················· | ···················· | ···················· |
| U | 394 | hCoV-19/England/MILK-B95E83/2020\|EPI_ISL_674987\|20 | ···················· | ···················· | ····················· | ···················· | ···················· | ···················· |
| U | 395 | hCoV-19/England/MILK-B971E2/2020\|EPI_ISL_674989\|20 | ···················· | ···················· | ····················· | ···················· | ···················· | ···················· |
| U | 396 | hCoV-19/England/MILK-B96DD0/2020\|EPI_ISL_674991\|20 | ···················· | ···················· | ····················· | ···················· | ···················· | ···················· |
| U | 397 | hCoV-19/England/CAMC-B98196/2020\|EPI_ISL_674992\|20 | ···················· | ···················· | ····················· | ···················· | ···················· | ···················· |
| U | 398 | hCoV-19/England/MILK-B96EFB/2020\|EPI_ISL_674993\|20 | ···················· | ·A·················· | ····················· | ···················· | ···················· | ···A················ |
| U | 399 | hCoV-19/England/MILK-B95ACB/2020\|EPI_ISL_674994\|20 | ···················· | ···················· | ····················· | ···················· | ···················· | ···················· |
| U | 400 | hCoV-19/England/MILK-B95C4D/2020\|EPI_ISL_674996\|20 | ···················· | ···················· | ····················· | ···················· | ···················· | ···················· |
| U | 401 | hCoV-19/England/MILK-B97267/2020\|EPI_ISL_674997\|20 | ···················· | ···················· | ····················· | ···················· | ···················· | ···················· |

| | | | | | | | | |
|---|---|---|---|---|---|---|---|---|
| U | 402 | hCoV-19/England/CAMC-B9862E/2020\|EPI_ISL_674998\|20 | ···················· | ···················· | ····················· | ···················· | ···················· | ···················· |
| U | 403 | hCoV-19/England/MILK-B97319/2020\|EPI_ISL_674999\|20 | ···················· | ·A·················· | ····················· | ···················· | ···················· | ···A················ |
| U | 404 | hCoV-19/USA/NY-Wadsworth-215499-01/2020\|EPI_ISL_67 | ···················· | ·A·················· | ····················· | ···················· | ···················· | ···················· |
| U | 405 | hCoV-19/USA/NY-Wadsworth-217641-01/2020\|EPI_ISL_67 | ···················· | ·A·················· | ····················· | ···················· | ···················· | ···················· |
| U | 406 | hCoV-19/USA/NY-Wadsworth-220230-01/2020\|EPI_ISL_67 | ···················· | ···················· | ····················· | ···················· | ···················· | ···················· |
| U | 407 | hCoV-19/USA/NY-Wadsworth-220320-01/2020\|EPI_ISL_67 | ···················· | ···················· | ····················· | ···················· | ···················· | ···················· |
| U | 408 | hCoV-19/USA/NY-Wadsworth-223644-01/2020\|EPI_ISL_67 | ···················· | ·A·················· | ····················· | ···················· | ···················· | ···················· |
| U | 409 | hCoV-19/USA/NY-Wadsworth-224392-01/2020\|EPI_ISL_67 | ···················· | ···················· | ····················· | ···················· | ···················· | ···················· |
| U | 410 | hCoV-19/USA/NY-Wadsworth-224393-01/2020\|EPI_ISL_67 | ···················· | ···················· | ····················· | ···················· | ···················· | ···················· |
| U | 411 | hCoV-19/USA/VA-DCLS-2064/2020\|EPI_ISL_677190\|2020- | ···················· | ···················· | ····················· | ···················· | ···················· | ···················· |
| U | 412 | hCoV-19/USA/VA-DCLS-2065/2020\|EPI_ISL_677191\|2020- | ···················· | ···················· | ····················· | ···················· | ···················· | ···················· |
| U | 413 | hCoV-19/USA/VA-DCLS-2066/2020\|EPI_ISL_677192\|2020- | ···················· | ···················· | ····················· | ···················· | ···················· | ···················· |
| U | 414 | hCoV-19/USA/VA-DCLS-2067/2020\|EPI_ISL_677193\|2020- | ···················· | ···················· | ····················· | ···················· | ···················· | ···················· |
| U | 415 | hCoV-19/USA/VA-DCLS-2165/2020\|EPI_ISL_677194\|2020- | ···················· | ···················· | ····················· | ···················· | ···················· | ···················· |
| U | 416 | hCoV-19/USA/VA-DCLS-2166/2020\|EPI_ISL_677195\|2020- | ···················· | ···················· | ····················· | ···················· | ···················· | ···················· |
| U | 417 | hCoV-19/USA/VA-DCLS-2167/2020\|EPI_ISL_677196\|2020- | ···················· | ···················· | ····················· | ···················· | ···················· | ···················· |
| U | 418 | hCoV-19/USA/VA-DCLS-2168/2020\|EPI_ISL_677197\|2020- | ···················· | ···················· | ····················· | ···················· | ···················· | ···················· |
| U | 419 | hCoV-19/USA/VA-DCLS-2169/2020\|EPI_ISL_677198\|2020- | ···················· | ···················· | ····················· | ···················· | ···················· | ···················· |
| U | 420 | hCoV-19/USA/VA-DCLS-2170/2020\|EPI_ISL_677199\|2020- | ···················· | ···················· | ····················· | ···················· | ···················· | ···················· |
| U | 421 | hCoV-19/USA/VA-DCLS-2058/2020\|EPI_ISL_677228\|2020- | ···················· | ···················· | ····················· | ···················· | ···················· | ···················· |
| U | 422 | hCoV-19/USA/VA-DCLS-2059/2020\|EPI_ISL_677229\|2020- | ···················· | ···················· | ····················· | ···················· | ···················· | ···················· |
| U | 423 | hCoV-19/USA/VA-DCLS-2060/2020\|EPI_ISL_677230\|2020- | ···················· | ···················· | ····················· | ···················· | ···················· | ···················· |
| U | 424 | hCoV-19/USA/VA-DCLS-2061/2020\|EPI_ISL_677231\|2020- | ···················· | ···················· | ····················· | ···················· | ···················· | ···················· |
| U | 425 | hCoV-19/USA/VA-DCLS-2062/2020\|EPI_ISL_677232\|2020- | ···················· | ···················· | ····················· | ···················· | ···················· | ···················· |
| U | 426 | hCoV-19/USA/VA-DCLS-2063/2020\|EPI_ISL_677233\|2020- | ···················· | ···················· | ····················· | ···················· | ···················· | ···················· |
| U | 427 | hCoV-19/USA/VA-DCLS-2113/2020\|EPI_ISL_677234\|2020- | ···················· | ···················· | ····················· | ···················· | ···················· | ···················· |
| U | 428 | hCoV-19/USA/VA-DCLS-2115/2020\|EPI_ISL_677235\|2020- | ···················· | ···················· | ····················· | ···················· | ···················· | ···················· |
| U | 429 | hCoV-19/USA/VA-DCLS-2116/2020\|EPI_ISL_677236\|2020- | ···················· | ·A·················· | ····················· | ···················· | ···················· | ···················· |
| U | 430 | hCoV-19/USA/VA-DCLS-2117/2020\|EPI_ISL_677237\|2020- | ···················· | ···················· | ····················· | ···················· | ···················· | ···················· |
| U | 431 | hCoV-19/USA/VA-DCLS-2118/2020\|EPI_ISL_677238\|2020- | ···················· | ·A·················· | ····················· | ···················· | ···················· | ···················· |
| U | 432 | hCoV-19/USA/VA-DCLS-2119/2020\|EPI_ISL_677239\|2020- | ···················· | ···················· | ····················· | ···················· | ···················· | ···················· |
| U | 433 | hCoV-19/USA/VA-DCLS-2120/2020\|EPI_ISL_677240\|2020- | ···················· | ·A·················· | ····················· | ···················· | ···················· | ···················· |
| U | 434 | hCoV-19/USA/CO-CDPHE-2009051621/2020\|EPI_ISL_67728 | ···················· | ···················· | ····················· | ···················· | ···················· | ···················· |
| U | 435 | hCoV-19/USA/CO-CDPHE-2003120912/2020\|EPI_ISL_67728 | ···················· | ···················· | ····················· | ···················· | ···················· | ···················· |
| U | 436 | hCoV-19/USA/CO-CDPHE-2003130078/2020\|EPI_ISL_67728 | ···················· | ···················· | ····················· | ···················· | ···················· | ···················· |
| U | 438 | hCoV-19/USA/CO-CDPHE-2004230036/2020\|EPI_ISL_67728 | ···················· | ···················· | ····················· | ···················· | ···················· | ···················· |
| U | 440 | hCoV-19/USA/CO-CDPHE-2005210130/2020\|EPI_ISL_67729 | ···················· | ···················· | ····················· | ···················· | ···················· | ···················· |
| U | 441 | hCoV-19/USA/CO-CDPHE-2005300277/2020\|EPI_ISL_67729 | ···················· | ···················· | ····················· | ···················· | ···················· | ···················· |
| U | 442 | hCoV-19/USA/CO-CDPHE-2006272103/2020\|EPI_ISL_67729 | ···················· | ···················· | ····················· | ···················· | ···················· | ···················· |
| U | 443 | hCoV-19/USA/CO-CDPHE-2007092984/2020\|EPI_ISL_67729 | ···················· | ···················· | ····················· | ···················· | ···················· | ···················· |
| U | 444 | hCoV-19/USA/CO-CDPHE-2007234429/2020\|EPI_ISL_67729 | ···················· | ···················· | ····················· | ···················· | ···················· | ···················· |
| U | 445 | hCoV-19/USA/CO-CDPHE-2007234441/2020\|EPI_ISL_67729 | ···················· | ···················· | ····················· | ···················· | ···················· | ···················· |
| U | 447 | hCoV-19/USA/CO-CDPHE-2008060288/2020\|EPI_ISL_67729 | ···················· | ·A·················· | ····················· | ···················· | ···················· | ···················· |
| U | 448 | hCoV-19/USA/CO-CDPHE-2008063119/2020\|EPI_ISL_67729 | ···················· | ···················· | ····················· | ···················· | ···················· | ···················· |
| U | 449 | hCoV-19/Belgium/ULG-10879/2020\|EPI_ISL_678422\|2020 | ···················· | ····A··············· | ····················· | ···················· | ···················· | ···················· |
| U | 450 | hCoV-19/Belgium/ULG-10884/2020\|EPI_ISL_678427\|2020 | ···················· | ···················· | ····················· | ···················· | ···················· | ···················· |
| U | 451 | hCoV-19/Belgium/ULG-10887/2020\|EPI_ISL_678430\|2020 | ···················· | ···················· | ····················· | ···················· | ···················· | ···················· |
| U | 452 | hCoV-19/Belgium/ULG-10889/2020\|EPI_ISL_678431\|2020 | ···················· | ···················· | ····················· | ···················· | ···················· | ···················· |
| U | 453 | hCoV-19/Belgium/ULG-10892/2020\|EPI_ISL_678434\|2020 | ···················· | ···················· | ····················· | ···················· | ···················· | ···················· |
| U | 454 | hCoV-19/Belgium/ULG-10893/2020\|EPI_ISL_678435\|2020 | ···················· | ···················· | ····················· | ···················· | ···················· | ···················· |
| U | 455 | hCoV-19/Belgium/ULG-10894/2020\|EPI_ISL_678436\|2020 | ···················· | ···················· | ····················· | ···················· | ···················· | ···················· |
| U | 456 | hCoV-19/Belgium/ULG-10895/2020\|EPI_ISL_678437\|2020 | ···················· | ···················· | ····················· | ···················· | ···················· | ···················· |
| U | 457 | hCoV-19/South Africa/KRISP-K005304/2020\|EPI_ISL_67 | ···················· | ···················· | ····················· | ···················· | ···················· | ···················· |
| U | 458 | hCoV-19/South Africa/KRISP-K005306/2020\|EPI_ISL_67 | ···················· | ···················· | ····················· | ···················· | ···················· | ···················· |
| U | 459 | hCoV-19/England/MILK-B3B2ED/2020\|EPI_ISL_679750\|20 | ···················· | ···················· | ····················· | ···················· | ···················· | ···················· |
| U | 460 | hCoV-19/England/MILK-B3B2FC/2020\|EPI_ISL_679751\|20 | ···················· | ···················· | ····················· | ···················· | ···················· | ···················· |
| U | 461 | hCoV-19/England/MILK-B3B308/2020\|EPI_ISL_679752\|20 | ···················· | ·A·················· | ····················· | ···················· | ···················· | ···A················ |
| U | 462 | hCoV-19/England/MILK-B3B317/2020\|EPI_ISL_679753\|20 | ···················· | ···················· | ····················· | ···················· | ···················· | ···················· |
| U | 463 | hCoV-19/England/MILK-B3B450/2020\|EPI_ISL_679758\|20 | ···················· | ···················· | ····················· | ···················· | ···················· | ···················· |
| U | 464 | hCoV-19/England/MILK-B3B46F/2020\|EPI_ISL_679759\|20 | ···················· | ···················· | ····················· | ···················· | ···················· | ···················· |
| U | 465 | hCoV-19/England/MILK-B3B511/2020\|EPI_ISL_679760\|20 | ···················· | ···················· | ····················· | ···················· | ···················· | ···················· |
| U | 466 | hCoV-19/England/MILK-B3B71B/2020\|EPI_ISL_679762\|20 | ···················· | ···················· | ····················· | ···················· | ···················· | ···················· |
| U | 467 | hCoV-19/England/MILK-B3B757/2020\|EPI_ISL_679766\|20 | ···················· | ···················· | ····················· | ···················· | ···················· | ···················· |
| U | 468 | hCoV-19/England/MILK-B3BC85/2020\|EPI_ISL_679800\|20 | ···················· | ···················· | ····················· | ···················· | ···················· | ···················· |
| U | 469 | hCoV-19/England/MILK-B3BC94/2020\|EPI_ISL_679801\|20 | ···················· | ···················· | ····················· | ···················· | ···················· | ···················· |
| U | 470 | hCoV-19/England/MILK-B3BD73/2020\|EPI_ISL_679808\|20 | ···················· | ···················· | ····················· | ···················· | ···················· | ···················· |
| U | 471 | hCoV-19/England/MILK-B3BD82/2020\|EPI_ISL_679809\|20 | ···················· | ···················· | ····················· | ···················· | ···················· | ···················· |
| U | 472 | hCoV-19/England/MILK-B599C6/2020\|EPI_ISL_679894\|20 | ···················· | ···················· | ····················· | ···················· | ···················· | ···················· |
| U | 473 | hCoV-19/England/MILK-B599D5/2020\|EPI_ISL_679895\|20 | ···················· | ···················· | ····················· | ···················· | ···················· | ···················· |
| U | 474 | hCoV-19/England/MILK-B599E4/2020\|EPI_ISL_679896\|20 | ···················· | ···················· | ····················· | ···················· | ···················· | ···················· |
| U | 475 | hCoV-19/England/MILK-B59A0F/2020\|EPI_ISL_679898\|20 | ···················· | ···················· | ····················· | ···················· | ···················· | ···················· |
| U | 476 | hCoV-19/Wales/PHWC-48DC30/2020\|EPI_ISL_680740\|2020 | ···················· | ···················· | ····················· | ···················· | ···················· | ···················· |
| U | 477 | hCoV-19/Wales/PHWC-48DC8B/2020\|EPI_ISL_680742\|2020 | ···················· | ······A············· | ····················· | ···················· | ···················· | ···················· |
| U | 478 | hCoV-19/Wales/PHWC-48DCA9/2020\|EPI_ISL_680743\|2020 | ···················· | ···················· | ····················· | ···················· | ···················· | ···················· |
| U | 479 | hCoV-19/Wales/PHWC-48DCC7/2020\|EPI_ISL_680745\|2020 | ···················· | ···················· | ····················· | ···················· | ···················· | ···················· |
| U | 480 | hCoV-19/Wales/PHWC-48E507/2020\|EPI_ISL_680870\|2020 | ···················· | ···················· | ····················· | ···················· | ···················· | ···················· |
| U | 483 | hCoV-19/Wales/PHWC-48E534/2020\|EPI_ISL_680873\|2020 | ···················· | ···················· | ····················· | ···················· | ···················· | ···················· |
| U | 484 | hCoV-19/Wales/PHWC-48E543/2020\|EPI_ISL_680874\|2020 | ···················· | ···················· | ····················· | ···················· | ···················· | ···················· |
| U | 485 | hCoV-19/Wales/PHWC-48E552/2020\|EPI_ISL_680875\|2020 | ···················· | ···················· | ····················· | ···················· | ···················· | ···················· |
| U | 487 | hCoV-19/Wales/PHWC-48E58F/2020\|EPI_ISL_680878\|2020 | ···················· | ···················· | ····················· | ···················· | ···················· | ···················· |

| | | | | | | | | |
|---|---|---|---|---|---|---|---|---|
| U | 489 hCoV-19/Wales/PHWC-48E5DA/2020\|EPI_ISL_680882\|2020 | ···················· | ···················· | ···················· | ···················· | ···················· | ···················· | |
| U | 491 hCoV-19/Wales/PHWC-48E5F8/2020\|EPI_ISL_680884\|2020 | ···················· | ···················· | ···················· | ···················· | ···················· | ···················· | |
| U | 493 hCoV-19/Wales/PHWC-48E622/2020\|EPI_ISL_680887\|2020 | ···················· | ···················· | ···················· | ···················· | ···················· | ···················· | |
| U | 495 hCoV-19/Wales/PHWC-48E640/2020\|EPI_ISL_680889\|2020 | ···················· | ···················· | ···················· | ···················· | ···················· | ···················· | |
| U | 496 hCoV-19/Wales/PHWC-48E68C/2020\|EPI_ISL_680892\|2020 | ···················· | ···················· | ···················· | ···················· | ···R················ | ···················· | |
| U | 497 hCoV-19/Wales/PHWC-48E69B/2020\|EPI_ISL_680893\|2020 | ···················· | ···················· | ···················· | ···················· | ···················· | ···················· | |
| U | 499 hCoV-19/Wales/PHWC-48E6C8/2020\|EPI_ISL_680896\|2020 | ···················· | ···················· | ···················· | ···················· | ···················· | ···················· | |
| U | 500 hCoV-19/Wales/PHWC-48E6D7/2020\|EPI_ISL_680897\|2020 | ···················· | ···················· | ···················· | ···················· | ···················· | ···················· | |
| U | 501 hCoV-19/Wales/PHWC-48E6E6/2020\|EPI_ISL_680898\|2020 | ···················· | ···················· | ···················· | ···················· | ···················· | ···················· | |
| U | 503 hCoV-19/Denmark/DCGC-12819/2020\|EPI_ISL_683000\|202 | ···················· | ···················· | ···················· | ···················· | ···················· | ···················· | |
| U | 504 hCoV-19/Denmark/DCGC-12822/2020\|EPI_ISL_683003\|202 | ···················· | ···················· | ···················· | ···················· | ···················· | ···················· | |
| U | 505 hCoV-19/Denmark/DCGC-12823/2020\|EPI_ISL_683004\|202 | ···················· | ····G··············· | ···················· | ···················· | ···················· | ···················· | |
| U | 506 hCoV-19/mink/Denmark/mDK-154/2020\|EPI_ISL_683005\|2 | ···················· | ···················· | ···················· | ···················· | ···················· | ···················· | |
| U | 507 hCoV-19/mink/Denmark/mDK-158/2020\|EPI_ISL_683009\|2 | ···················· | ···················· | ···················· | ···················· | ···················· | ···················· | |
| U | 508 hCoV-19/mink/Denmark/mDK-159/2020\|EPI_ISL_683010\|2 | ···················· | ···················· | ···················· | ···················· | ···················· | ···················· | |
| U | 509 hCoV-19/mink/Denmark/mDK-172/2020\|EPI_ISL_683023\|2 | ···················· | ···················· | ···················· | ···················· | ···················· | ···················· | |
| U | 510 hCoV-19/mink/Denmark/mDK-173/2020\|EPI_ISL_683024\|2 | ···················· | ···················· | ···················· | ···················· | ···················· | ···················· | |
| U | 511 hCoV-19/mink/Denmark/mDK-174/2020\|EPI_ISL_683025\|2 | ···················· | ···················· | ···················· | ···················· | ···················· | ···················· | |
| U | 512 hCoV-19/mink/Denmark/mDK-190/2020\|EPI_ISL_683041\|2 | ···················· | ···················· | ···················· | ···················· | ···················· | ···················· | |
| U | 513 hCoV-19/England/MILK-B97355/2020\|EPI_ISL_674901\|20 | ···················· | ···················· | ···················· | ···················· | ···················· | ···················· | |
| U | 514 hCoV-19/England/MILK-B96471/2020\|EPI_ISL_674904\|20 | ···················· | ···················· | ···················· | ···················· | ···················· | ···················· | |
| U | 515 hCoV-19/England/MILK-B95A9E/2020\|EPI_ISL_674905\|20 | ···················· | ···················· | ···················· | ···················· | ···················· | ···················· | |
| U | 516 hCoV-19/England/CAMC-B9822A/2020\|EPI_ISL_674906\|20 | ···················· | ···················· | ···················· | ···················· | ···················· | ···················· | |
| U | 517 hCoV-19/England/MILK-B975B9/2020\|EPI_ISL_674907\|20 | ···················· | ···················· | ···················· | ···················· | ···················· | ···················· | |
| U | 518 hCoV-19/England/CAMC-B9814B/2020\|EPI_ISL_674908\|20 | ···················· | ···················· | ···················· | ···················· | ···················· | ···················· | |
| U | 519 hCoV-19/England/CAMC-B9815A/2020\|EPI_ISL_674909\|20 | ···················· | ···················· | ···················· | ···················· | ···················· | ···················· | |
| U | 520 hCoV-19/USA/NY-Wadsworth-226490-01/2020\|EPI_ISL_67 | ···················· | ···················· | ···················· | ···················· | ···················· | ···················· | |
| U | 521 hCoV-19/USA/NY-Wadsworth-226491-01/2020\|EPI_ISL_67 | ···················· | ···················· | ···················· | ···················· | ···················· | ···················· | |
| U | 522 hCoV-19/USA/NY-Wadsworth-226492-01/2020\|EPI_ISL_67 | ···················· | ···················· | ···················· | ···················· | ···················· | ···················· | |
| U | 523 hCoV-19/USA/NY-Wadsworth-226513-01/2020\|EPI_ISL_67 | ···················· | ···················· | ···················· | ···················· | ···················· | ···················· | |
| U | 524 hCoV-19/USA/NY-Wadsworth-226514-01/2020\|EPI_ISL_67 | ···················· | ···················· | ···················· | ···················· | ···················· | ···················· | |
| U | 525 hCoV-19/USA/NY-Wadsworth-226515-01/2020\|EPI_ISL_67 | ···················· | ···················· | ···················· | ···················· | ···················· | ···················· | |
| U | 526 hCoV-19/USA/NY-Wadsworth-226516-01/2020\|EPI_ISL_67 | ···················· | ···················· | ···················· | ···················· | ···················· | ···················· | |
| U | 527 hCoV-19/USA/NY-Wadsworth-226517-01/2020\|EPI_ISL_67 | ···················· | ···················· | ···················· | ···················· | ···················· | ···················· | |
| U | 528 hCoV-19/USA/NY-Wadsworth-226519-01/2020\|EPI_ISL_67 | ···················· | ···················A | ···················· | ···················· | ···················· | ···················· | 1 |
| U | 529 hCoV-19/USA/NY-Wadsworth-226966-01/2020\|EPI_ISL_67 | ···················· | ···················· | ···················· | ···················· | ···················· | ···················· | |
| U | 530 hCoV-19/USA/NY-Wadsworth-228415-01/2020\|EPI_ISL_67 | ···················· | ···················· | ···················· | ···················· | ···················· | ···················· | |
| U | 531 hCoV-19/USA/NY-Wadsworth-229167-01/2020\|EPI_ISL_67 | ···················· | ···················· | ···················· | ···················· | ···················· | ···················· | |
| U | 532 hCoV-19/USA/NY-Wadsworth-232102-01/2020\|EPI_ISL_67 | ···················· | ···················· | ···················· | ···················· | ···················· | ···················· | |
| U | 533 hCoV-19/USA/VA-DCLS-2091/2020\|EPI_ISL_677134\|2020- | ···················· | ···················· | ···················· | ···················· | ···················· | ···················· | |
| U | 534 hCoV-19/USA/VA-DCLS-2093/2020\|EPI_ISL_677135\|2020- | ···················· | ···················· | ···················· | ···················· | ···················· | ···················· | |
| U | 535 hCoV-19/USA/VA-DCLS-2094/2020\|EPI_ISL_677136\|2020- | ···················· | ···················· | ···················· | ···················· | ···················· | ···················· | |
| U | 536 hCoV-19/USA/VA-DCLS-2095/2020\|EPI_ISL_677137\|2020- | ···················· | ···················· | ···················· | ···················· | ···················· | ···················· | |
| U | 537 hCoV-19/USA/VA-DCLS-2100/2020\|EPI_ISL_677138\|2020- | ···················· | ···················· | ···················· | ···················· | ···················· | ···················· | |
| U | 538 hCoV-19/USA/VA-DCLS-2101/2020\|EPI_ISL_677139\|2020- | ···················· | ···················· | ···················· | ···················· | ···················· | ···················· | |
| U | 539 hCoV-19/USA/VA-DCLS-2104/2020\|EPI_ISL_677140\|2020- | ···················· | ···················· | ···················· | ···················· | ···················· | ···················· | |
| U | 540 hCoV-19/USA/VA-DCLS-2106/2020\|EPI_ISL_677141\|2020- | ···················· | ···················· | ···················· | ···················· | ···················· | ···················· | |
| U | 541 hCoV-19/USA/VA-DCLS-2108/2020\|EPI_ISL_677142\|2020- | ···················· | ···················· | ···················· | ···················· | ···················· | ···················· | |
| U | 542 hCoV-19/USA/VA-DCLS-2109/2020\|EPI_ISL_677143\|2020- | ···················· | ···················· | ···················· | ···················· | ···················· | ···················· | |
| U | 543 hCoV-19/USA/VA-DCLS-2110/2020\|EPI_ISL_677144\|2020- | ···················· | ·A·················· | ···················· | ···················· | ···················· | ···················· | |
| U | 544 hCoV-19/USA/VA-DCLS-2111/2020\|EPI_ISL_677145\|2020- | ···················· | ·A·················· | ···················· | ···················· | ···················· | ···················· | |
| U | 545 hCoV-19/USA/VA-DCLS-2112/2020\|EPI_ISL_677146\|2020- | ···················· | ·····C·············· | ···················· | ···················· | ···················· | ···················· | |
| U | 546 hCoV-19/USA/VA-DCLS-2122/2020\|EPI_ISL_677147\|2020- | ···················· | ···················· | ···················· | ···················· | ···················· | ···················· | |
| U | 547 hCoV-19/USA/VA-DCLS-2123/2020\|EPI_ISL_677148\|2020- | ···················· | ···················· | ···················· | ···················· | ···················· | ···················· | |
| U | 548 hCoV-19/USA/VA-DCLS-2124/2020\|EPI_ISL_677149\|2020- | ···················· | ···················· | ···················· | ···················· | ···················· | ···················· | |
| U | 549 hCoV-19/USA/VA-DCLS-2125/2020\|EPI_ISL_677150\|2020- | ···················· | ···················· | ···················· | ···················· | ···················· | ···················· | |
| U | 550 hCoV-19/USA/VA-DCLS-2126/2020\|EPI_ISL_677151\|2020- | ···················· | ···················· | ···················· | ···················· | ···················· | ···················· | |
| U | 551 hCoV-19/USA/VA-DCLS-2128/2020\|EPI_ISL_677153\|2020- | ···················· | ···················· | ···················· | ···················· | ···················· | ···················· | |
| U | 552 hCoV-19/USA/VA-DCLS-2129/2020\|EPI_ISL_677154\|2020- | ···················· | ···················· | ···················· | ···················· | ···················· | ···················· | |
| U | 553 hCoV-19/USA/VA-DCLS-2131/2020\|EPI_ISL_677155\|2020- | ···················· | ···················· | ···················· | ···················· | ···················· | ···················· | |
| U | 554 hCoV-19/USA/VA-DCLS-2133/2020\|EPI_ISL_677156\|2020- | ···················· | ···················· | ···················· | ···················· | ···················· | ···················· | |
| U | 555 hCoV-19/USA/VA-DCLS-2135/2020\|EPI_ISL_677157\|2020- | ···················· | ·A·················· | ···················· | ···················· | ···················· | ···················· | |
| U | 556 hCoV-19/USA/VA-DCLS-2136/2020\|EPI_ISL_677158\|2020- | ···················· | ···················· | ···················· | ···················· | ···················· | ···················· | |
| U | 557 hCoV-19/USA/VA-DCLS-2137/2020\|EPI_ISL_677159\|2020- | ···················· | ·A·················· | ···················· | ···················· | ···················· | ···················· | |
| U | 558 hCoV-19/USA/VA-DCLS-2139/2020\|EPI_ISL_677160\|2020- | ···················· | ·A·················· | ···················· | ···················· | ···················· | ···················· | |
| U | 559 hCoV-19/USA/VA-DCLS-2142/2020\|EPI_ISL_677161\|2020- | ···················· | ···················· | ···················· | ···················· | ···················· | ···················· | |
| U | 560 hCoV-19/USA/VA-DCLS-2143/2020\|EPI_ISL_677162\|2020- | ···················· | ···················· | ···················· | ···················· | ···················· | ···················· | |
| U | 561 hCoV-19/USA/VA-DCLS-2144/2020\|EPI_ISL_677163\|2020- | ···················· | ···················· | ···················· | ···················· | ···················· | ···················· | |
| U | 562 hCoV-19/USA/VA-DCLS-2011/2020\|EPI_ISL_677164\|2020- | ···················· | ·A·················· | ···················· | ···················· | ···················· | ···················· | |
| U | 563 hCoV-19/USA/VA-DCLS-2012/2020\|EPI_ISL_677165\|2020- | ···················· | ···················· | ···················· | ···················· | ···················· | ···················· | |
| U | 564 hCoV-19/USA/VA-DCLS-2015/2020\|EPI_ISL_677166\|2020- | ···················· | ·A·················· | ···················· | ···················· | ···················· | ···················· | |
| U | 565 hCoV-19/USA/VA-DCLS-2018/2020\|EPI_ISL_677167\|2020- | ···················· | ···················· | ···················· | ···················· | ···················· | ···················· | |
| U | 566 hCoV-19/USA/VA-DCLS-2019/2020\|EPI_ISL_677168\|2020- | ···················· | ·A·················· | ···················· | ···················· | ···················· | ···················· | |
| U | 567 hCoV-19/USA/VA-DCLS-2021/2020\|EPI_ISL_677169\|2020- | ···················· | ···················· | ···················· | ···················· | ···················· | ···················· | |
| U | 568 hCoV-19/USA/VA-DCLS-2022/2020\|EPI_ISL_677170\|2020- | ···················· | ···················· | ···················· | ···················· | ···················· | ···················· | |
| U | 569 hCoV-19/USA/VA-DCLS-2025/2020\|EPI_ISL_677171\|2020- | ···················· | ···················· | ···················· | ···················· | ···················· | ···················· | |
| U | 570 hCoV-19/USA/VA-DCLS-2027/2020\|EPI_ISL_677172\|2020- | ···················· | ···················· | ···················· | ···················· | ···················· | ···················· | |
| U | 571 hCoV-19/USA/VA-DCLS-2028/2020\|EPI_ISL_677173\|2020- | ···················· | ···················· | ···················· | ···················· | ···················· | ···················· | |
| U | 572 hCoV-19/USA/VA-DCLS-2030/2020\|EPI_ISL_677174\|2020- | ···················· | ···················· | ···················· | ···················· | ···················· | ···················· | |
| U | 573 hCoV-19/USA/VA-DCLS-2031/2020\|EPI_ISL_677175\|2020- | ···················· | ·A·················· | ···················· | ···················· | ···················· | ···················· | |

| | | | | | | | |
|---|---|---|---|---|---|---|---|
| U | 574 hCoV-19/USA/VA-DCLS-2032/2020\|EPI_ISL_677176\|2020- | ···················· | ···················· | ····················· | ···················· | ···················· | ···················· |
| U | 575 hCoV-19/USA/VA-DCLS-2033/2020\|EPI_ISL_677177\|2020- | ···················· | ···················· | ····················· | ···················· | ···················· | ···················· |
| U | 576 hCoV-19/USA/VA-DCLS-2036/2020\|EPI_ISL_677178\|2020- | ···················· | ···················· | ····················· | ···················· | ···················· | ···················· |
| U | 577 hCoV-19/USA/VA-DCLS-2069/2020\|EPI_ISL_677179\|2020- | ···················· | ····A··············· | ····················· | ···················· | ···················· | ···················· |
| U | 578 hCoV-19/USA/VA-DCLS-2072/2020\|EPI_ISL_677180\|2020- | ···················· | ···················· | ····················· | ···················· | ···················· | ···················· |
| U | 579 hCoV-19/USA/VA-DCLS-2073/2020\|EPI_ISL_677181\|2020- | ···················· | ···················· | ····················· | ···················· | ···················· | ···················· |
| U | 580 hCoV-19/USA/VA-DCLS-2075/2020\|EPI_ISL_677183\|2020- | ···················· | ···················· | ····················· | ···················· | ···················· | ···················· |
| U | 581 hCoV-19/USA/VA-DCLS-2076/2020\|EPI_ISL_677184\|2020- | ···················· | ···················· | ····················· | ···················· | ···················· | ···················· |
| U | 582 hCoV-19/USA/VA-DCLS-2078/2020\|EPI_ISL_677185\|2020- | ···················· | ···················· | ····················· | ···················· | ···················· | ···················· |
| U | 583 hCoV-19/USA/VA-DCLS-2079/2020\|EPI_ISL_677186\|2020- | ···················· | ···················· | ····················· | ···················· | ···················· | ···················· |
| U | 584 hCoV-19/USA/VA-DCLS-2080/2020\|EPI_ISL_677187\|2020- | ···················· | ···················· | ····················· | ···················· | ···················· | ···················· |
| U | 585 hCoV-19/USA/VA-DCLS-2084/2020\|EPI_ISL_677188\|2020- | ···················· | ···················· | ····················· | ···················· | ···················· | ···················· |
| U | 586 hCoV-19/USA/VA-DCLS-2089/2020\|EPI_ISL_677189\|2020- | ···················· | ···················· | ····················· | ···················· | ···················· | ···················· |
| U | 587 hCoV-19/Belgium/ULG-10856/2020\|EPI_ISL_678400\|2020 | ···················· | ···················· | ····················· | ···················· | ···················· | ···················· |
| U | 588 hCoV-19/Belgium/ULG-10857/2020\|EPI_ISL_678401\|2020 | ·T·················· | ···················· | ····················· | ···················· | ···················· | ···················· |
| U | 589 hCoV-19/Belgium/ULG-10858/2020\|EPI_ISL_678402\|2020 | ···················· | ···················· | ····················· | ···················· | ···················· | ···················· |
| U | 590 hCoV-19/Belgium/ULG-10859/2020\|EPI_ISL_678403\|2020 | ···················· | ···················· | ····················· | ···················· | ···················· | ···················· |
| U | 591 hCoV-19/Belgium/ULG-10860/2020\|EPI_ISL_678404\|2020 | ···················· | ···················· | ····················· | ···················· | ···················· | ···················· |
| U | 592 hCoV-19/Belgium/ULG-10861/2020\|EPI_ISL_678405\|2020 | ···················· | ···················· | ····················· | ···················· | ···················· | ···················· |
| U | 593 hCoV-19/Belgium/ULG-10862/2020\|EPI_ISL_678406\|2020 | ···················· | ···················· | ····················· | ···················· | ···················· | ···················· |
| U | 594 hCoV-19/Belgium/ULG-10863/2020\|EPI_ISL_678407\|2020 | ···················· | ·A·················· | ····················· | ···················· | ···················· | ···················· |
| U | 595 hCoV-19/Belgium/ULG-10864/2020\|EPI_ISL_678408\|2020 | ···················· | ···················· | ····················· | ···················· | ···················· | ···················· |
| U | 596 hCoV-19/Belgium/ULG-10867/2020\|EPI_ISL_678411\|2020 | ···················· | ···················· | ····················· | ···················· | ···················· | ···················· |
| U | 597 hCoV-19/Belgium/ULG-10874/2020\|EPI_ISL_678417\|2020 | ···················· | ···················· | ····················· | ···················· | ···················· | ···················· |
| U | 598 hCoV-19/Belgium/ULG-10875/2020\|EPI_ISL_678418\|2020 | ···················· | ···················· | ····················· | ···················· | ···················· | ···················· |
| U | 599 hCoV-19/Belgium/ULG-10882/2020\|EPI_ISL_678425\|2020 | ···················· | ···················· | ····················· | ···················· | ···················· | ···················· |
| U | 600 hCoV-19/Belgium/ULG-10883/2020\|EPI_ISL_678426\|2020 | ···················· | ···················· | ····················· | ···················· | ···················· | ···················· |
| U | 601 hCoV-19/Belgium/ULG-10912/2020\|EPI_ISL_678453\|2020 | ···················· | ···················· | ····················· | ···················· | ······T············· | ···················· |
| U | 602 hCoV-19/Belgium/ULG-10915/2020\|EPI_ISL_678456\|2020 | ···················· | ···················· | ····················· | ···················· | ···················· | ···················· |
| U | 603 hCoV-19/Belgium/ULG-10916/2020\|EPI_ISL_678457\|2020 | ···················· | ···················· | ····················· | ···················· | ···················· | ···················· |
| U | 604 hCoV-19/Belgium/ULG-10918/2020\|EPI_ISL_678459\|2020 | ···················· | ···················· | ····················· | ···················· | ······T············· | ···················· |
| U | 605 hCoV-19/Belgium/ULG-10920/2020\|EPI_ISL_678460\|2020 | ···················· | ···················· | ····················· | ···················· | ···················· | ···················· |
| U | 606 hCoV-19/Belgium/ULG-10921/2020\|EPI_ISL_678461\|2020 | ···················· | ···················· | ····················· | ···················· | ···················· | ···················· |
| U | 607 hCoV-19/Belgium/ULG-10924/2020\|EPI_ISL_678464\|2020 | ···················· | ···················· | ····················· | ···················· | ···················· | ···················· |
| U | 608 hCoV-19/Belgium/ULG-10925/2020\|EPI_ISL_678465\|2020 | ···················· | ···················· | ····················· | ···················· | ···················· | ···················· |
| U | 609 hCoV-19/Belgium/ULG-10926/2020\|EPI_ISL_678466\|2020 | ···················· | ···················· | ····················· | ···················· | ···················· | ···················· |
| U | 610 hCoV-19/Belgium/ULG-10928/2020\|EPI_ISL_678468\|2020 | ···················· | ···················· | ····················· | ···················· | ···················· | ···················· |
| U | 611 hCoV-19/Belgium/ULG-10930/2020\|EPI_ISL_678469\|2020 | ···················· | ···················· | ····················· | ···················· | ···················· | ···················· |
| U | 612 hCoV-19/Belgium/ULG-10931/2020\|EPI_ISL_678470\|2020 | ···················· | ···················· | ····················· | ···················· | ···················· | ···················· |
| U | 613 hCoV-19/Belgium/ULG-10933/2020\|EPI_ISL_678472\|2020 | ···················· | ···················· | ····················· | ···················· | ···················· | ···················· |
| U | 614 hCoV-19/Belgium/ULG-10934/2020\|EPI_ISL_678473\|2020 | ···················· | ···················· | ····················· | ···················· | ···················· | ···················· |
| U | 615 hCoV-19/Belgium/ULG-10935/2020\|EPI_ISL_678474\|2020 | ···················· | ···················· | ·········T··········· | ···················· | ···················· | ···················· |
| U | 616 hCoV-19/Belgium/ULG-10936/2020\|EPI_ISL_678475\|2020 | ···················· | ···················· | ····················· | ···················· | ···················· | ···················· |
| U | 617 hCoV-19/Belgium/ULG-10937/2020\|EPI_ISL_678476\|2020 | ···················· | ···················· | ····················· | ···················· | ···················· | ···················· |
| U | 618 hCoV-19/Belgium/ULG-10938/2020\|EPI_ISL_678477\|2020 | ···················· | ···················· | ····················· | ···················· | ···················· | ···················· |
| U | 619 hCoV-19/Belgium/ULG-10939/2020\|EPI_ISL_678478\|2020 | ···················· | ···················· | ····················· | ···················· | ···················· | ···················· |
| U | 620 hCoV-19/Belgium/ULG-10940/2020\|EPI_ISL_678479\|2020 | ···················· | ···················· | ····················· | ···················· | ······T············· | ···················· |
| U | 621 hCoV-19/Belgium/ULG-10941/2020\|EPI_ISL_678480\|2020 | ···················· | ···················· | ····················· | ···················· | ···················· | ···················· |
| U | 622 hCoV-19/Belgium/ULG-10944/2020\|EPI_ISL_678483\|2020 | ···················· | ···················· | ····················· | ···················· | ···················· | ···················· |
| U | 623 hCoV-19/Belgium/ULG-10945/2020\|EPI_ISL_678484\|2020 | ···················· | ···················· | ····················· | ···················· | ···················· | ···················· |
| U | 624 hCoV-19/Belgium/ULG-10946/2020\|EPI_ISL_678485\|2020 | ···················· | ···················· | ····················· | ···················· | ···················· | ···················· |
| U | 625 hCoV-19/Serbia/Sabac-000012108/2020\|EPI_ISL_678486 | ···················· | ···················· | ····················· | ···················· | ···················· | ···················· |
| U | 626 hCoV-19/Serbia/Sabac-000282104/2020\|EPI_ISL_678487 | ···················· | ···················· | ····················· | ···················· | ···················· | ···················· |
| U | 627 hCoV-19/Serbia/Sabac-000533004/2020\|EPI_ISL_678488 | ···················· | ···················· | ····················· | ···················· | ···················· | ···················· |
| U | 628 hCoV-19/Serbia/Sabac-000020508/2020\|EPI_ISL_678489 | ···················· | ···················· | ····················· | ···················· | ···················· | ···················· |
| U | 629 hCoV-19/Serbia/Sabac-000573004/2020\|EPI_ISL_678490 | ···················· | ···················· | ····················· | ···················· | ······T············· | ···················· |
| U | 630 hCoV-19/Serbia/Sabac-000230907/2020\|EPI_ISL_678491 | ···················· | ···················· | ····················· | ···················· | ···················· | ···················· |
| U | 631 hCoV-19/Serbia/Sabac-000490807/2020\|EPI_ISL_678492 | ···················· | ···················· | ····················· | ···················· | ···················· | ···················· |
| U | 632 hCoV-19/Serbia/Sabac-001701607/2020\|EPI_ISL_678493 | ···················· | ···················· | ····················· | ···················· | ···················· | ···················· |
| U | 633 hCoV-19/France/ARA-9315/2020\|EPI_ISL_678494\|2020-0 | ···················· | ···················· | ····················· | ···················· | ···················· | ···················· |
| U | 634 hCoV-19/France/ARA-9566/2020\|EPI_ISL_678495\|2020-0 | ···················· | ···················· | ····················· | ···················· | ···················· | ···················· |
| U | 635 hCoV-19/France/ARA-12561/2020\|EPI_ISL_678496\|2020- | ···················· | ···················· | ····················· | ···················· | ···················· | ···················· |
| U | 636 hCoV-19/France/ARA-14177/2020\|EPI_ISL_678497\|2020- | ···················· | ···················· | ····················· | ···················· | ···················· | ···················· |
| U | 637 hCoV-19/France/ARA-15456/2020\|EPI_ISL_678498\|2020- | ···················· | ···················· | ····················· | ···················· | ···················· | ···················· |
| U | 638 hCoV-19/France/ARA-15829/2020\|EPI_ISL_678499\|2020- | ···················· | ···················· | ····················· | ···················· | ···················· | ···················· |
| U | 639 hCoV-19/England/MILK-B3A9E9/2020\|EPI_ISL_679700\|20 | ···················· | ···················· | ····················· | ···················· | ···················· | ···················· |
| U | 640 hCoV-19/England/MILK-B3AA04/2020\|EPI_ISL_679701\|20 | ···················· | ···················· | ····················· | ···················· | ···················· | ···················· |
| U | 641 hCoV-19/England/MILK-B3AA13/2020\|EPI_ISL_679702\|20 | ···················· | ···················· | ····················· | ···················· | ···················· | ···················· |
| U | 642 hCoV-19/England/MILK-B3AA31/2020\|EPI_ISL_679703\|20 | ···················· | ···················· | ····················· | ···················· | ···················· | ···················· |
| U | 643 hCoV-19/England/MILK-B3AA40/2020\|EPI_ISL_679704\|20 | ···················· | ···················· | ····················· | ···················· | ···················· | ···A················ |
| U | 644 hCoV-19/England/MILK-B3AB2F/2020\|EPI_ISL_679705\|20 | ···················· | ···················· | ····················· | ···················· | ···················· | ···················· |
| U | 645 hCoV-19/England/MILK-B3AB3E/2020\|EPI_ISL_679706\|20 | ···················· | ···················· | ····················· | ···················· | ···················· | ···················· |
| U | 646 hCoV-19/England/MILK-B3AB5C/2020\|EPI_ISL_679707\|20 | ···················· | ···················· | ····················· | ···················· | ···················· | ···················· |
| U | 647 hCoV-19/England/MILK-B3AB6B/2020\|EPI_ISL_679708\|20 | ···················· | ···················· | ····················· | ···················· | ···················· | ···················· |
| U | 648 hCoV-19/England/MILK-B3AB89/2020\|EPI_ISL_679709\|20 | ···················· | ···················· | ····················· | ···················· | ···················· | ···················· |
| U | 649 hCoV-19/England/MILK-B3AB98/2020\|EPI_ISL_679710\|20 | ···················· | ···················· | ····················· | ···················· | ···················· | ···················· |
| U | 650 hCoV-19/England/MILK-B3ABA7/2020\|EPI_ISL_679711\|20 | ···················· | ···················· | ····················· | ···················· | ···················· | ···················· |
| U | 651 hCoV-19/England/MILK-B3ABB6/2020\|EPI_ISL_679712\|20 | ···················· | ···················· | ····················· | ···················· | ···················· | ···················· |
| U | 652 hCoV-19/England/MILK-B3ABC5/2020\|EPI_ISL_679713\|20 | ···················· | ···················· | ····················· | ···················· | ···················· | ···················· |
| U | 653 hCoV-19/England/MILK-B3ABD4/2020\|EPI_ISL_679714\|20 | ···················· | ···················· | ····················· | ···················· | ···················· | ···················· |

| | | | | | | | |
|---|---|---|---|---|---|---|---|
| U | 654 hCoV-19/England/MILK-B3ABF2/2020\|EPI_ISL_679715\|20 | ···················· | ···················· | ···················· | ···················· | ···················· | ···················· |
| U | 655 hCoV-19/England/MILK-B3AC1D/2020\|EPI_ISL_679716\|20 | ···················· | ···················· | ···················· | ···················· | ···················· | ···················· |
| U | 656 hCoV-19/England/MILK-B3AC2C/2020\|EPI_ISL_679717\|20 | ···················· | ···················· | ···················· | ···················· | ···················· | ···················· |
| U | 657 hCoV-19/England/MILK-B3AC3B/2020\|EPI_ISL_679718\|20 | ···················· | ···················· | ···················· | ···················· | ···················· | ···················· |
| U | 658 hCoV-19/England/MILK-B3AC4A/2020\|EPI_ISL_679719\|20 | ···················· | ···················· | ···················· | ···················· | ···················· | ···················· |
| U | 659 hCoV-19/England/MILK-B3AC59/2020\|EPI_ISL_679720\|20 | ···················· | ···················· | ···················· | ···················· | ···················· | ···················· |
| U | 660 hCoV-19/England/MILK-B3AC68/2020\|EPI_ISL_679721\|20 | ···················· | ···················· | ···················· | ···················· | ······T············· | ···················· |
| U | 661 hCoV-19/England/MILK-B3ACB3/2020\|EPI_ISL_679722\|20 | ···················· | ···················· | ···················· | ···················· | ···················· | ···················· |
| U | 662 hCoV-19/England/MILK-B3AD0B/2020\|EPI_ISL_679724\|20 | ···················· | ···················· | ···················· | ···················· | ···················· | ···················· |
| U | 663 hCoV-19/England/MILK-B3AD29/2020\|EPI_ISL_679725\|20 | ···················· | ···················· | ···················· | ···················· | ···················· | ···················· |
| U | 664 hCoV-19/England/MILK-B3AD38/2020\|EPI_ISL_679726\|20 | ···················· | ···················· | ···················· | ···················· | ···················· | ···················· |
| U | 665 hCoV-19/England/MILK-B3AD65/2020\|EPI_ISL_679727\|20 | ···················· | ·A·················· | ···················· | ···················· | ···················· | ···················· |
| U | 666 hCoV-19/England/MILK-B3AD74/2020\|EPI_ISL_679728\|20 | ···················· | ·A·················· | ···················· | ···················· | ···················· | ···················· |
| U | 667 hCoV-19/England/MILK-B3AD83/2020\|EPI_ISL_679729\|20 | ···················· | ···················· | ···················· | ···················· | ···················· | ···················· |
| U | 668 hCoV-19/England/MILK-B3ADB0/2020\|EPI_ISL_679730\|20 | ···················· | ···················· | ···················· | ···················· | ···················· | ···················· |
| U | 669 hCoV-19/England/MILK-B3ADED/2020\|EPI_ISL_679731\|20 | ···················· | ···················· | ···················· | ···················· | ······T············· | ···················· |
| U | 670 hCoV-19/England/MILK-B3ADFC/2020\|EPI_ISL_679732\|20 | ···················· | ···················· | ···················· | ···················· | ···················· | ···················· |
| U | 671 hCoV-19/England/MILK-B3AFD8/2020\|EPI_ISL_679733\|20 | ···················· | ···················· | ···················· | ···················· | ···················· | ···················· |
| U | 672 hCoV-19/England/MILK-B3AFF6/2020\|EPI_ISL_679734\|20 | ···················· | ···················· | ···················· | ···················· | ···················· | ···················· |
| U | 673 hCoV-19/England/MILK-B3B010/2020\|EPI_ISL_679735\|20 | ···················· | ···················· | ···················· | ···················· | ···················· | ···················· |
| U | 674 hCoV-19/England/MILK-B3B02F/2020\|EPI_ISL_679736\|20 | ···················· | ···················· | ···················· | ···················· | ···················· | ···················· |
| U | 675 hCoV-19/England/MILK-B3B04D/2020\|EPI_ISL_679737\|20 | ···················· | ···················· | ···················· | ···················· | ···················· | ···················· |
| U | 676 hCoV-19/England/MILK-B3B06B/2020\|EPI_ISL_679738\|20 | ···················· | ·A·················· | ···················· | ···················· | ···················· | ···A················ |
| U | 677 hCoV-19/England/MILK-B3B07A/2020\|EPI_ISL_679739\|20 | ···················· | ···················· | ···················· | ···················· | ···················· | ···················· |
| U | 678 hCoV-19/England/MILK-B3B089/2020\|EPI_ISL_679740\|20 | ···················· | ···················· | ···················· | ···················· | ···················· | ···················· |
| U | 679 hCoV-19/England/MILK-B3B0C5/2020\|EPI_ISL_679741\|20 | ···················· | ···················· | ···················· | ···················· | ···················· | ···················· |
| U | 680 hCoV-19/England/MILK-B3B0E3/2020\|EPI_ISL_679742\|20 | ···················· | ···················· | ···················· | ···················· | ···················· | ···················· |
| U | 681 hCoV-19/England/MILK-B3B0F2/2020\|EPI_ISL_679743\|20 | ···················· | ···················· | ···················· | ···················· | ···················· | ···················· |
| U | 682 hCoV-19/England/MILK-B3B10E/2020\|EPI_ISL_679744\|20 | ···················· | ···················· | ···················· | ···················· | ···················· | ···················· |
| U | 683 hCoV-19/England/MILK-B3B11D/2020\|EPI_ISL_679745\|20 | ···················· | ···················· | ···················· | ···················· | ···················· | ···················· |
| U | 684 hCoV-19/England/MILK-B3B2A1/2020\|EPI_ISL_679746\|20 | ···················· | ···················· | ···················· | ···················· | ···················· | ···················· |
| U | 685 hCoV-19/England/MILK-B3B2B0/2020\|EPI_ISL_679747\|20 | ···················· | ···················· | ···················· | ···················· | ···················· | ···················· |
| U | 686 hCoV-19/England/MILK-B3B326/2020\|EPI_ISL_679754\|20 | ···················· | ···················· | ···················· | ···················· | ···················· | ···················· |
| U | 687 hCoV-19/England/MILK-B3B335/2020\|EPI_ISL_679755\|20 | ···················· | ···················· | ···················· | ···················· | ···················· | ···················· |
| U | 688 hCoV-19/England/MILK-B3B414/2020\|EPI_ISL_679756\|20 | ···················· | ···················· | ···················· | ···················· | ···················· | ···················· |
| U | 689 hCoV-19/England/MILK-B3B432/2020\|EPI_ISL_679757\|20 | ···················· | ·A·················· | ···················· | ···················· | ···················· | ···A················ |
| U | 690 hCoV-19/England/MILK-B3B872/2020\|EPI_ISL_679780\|20 | ···················· | ···················· | ···················· | ···················· | ···················· | ···················· |
| U | 691 hCoV-19/England/MILK-B3B97F/2020\|EPI_ISL_679781\|20 | ···················· | ···················· | ···················· | ···················· | ······T············· | ···················· |
| U | 692 hCoV-19/England/MILK-B3B9BB/2020\|EPI_ISL_679782\|20 | ···················· | ···················· | ···················· | ···················· | ···················· | ···················· |
| U | 693 hCoV-19/England/MILK-B3B9D9/2020\|EPI_ISL_679783\|20 | ···················· | ···················· | ···················· | ···················· | ···················· | ···················· |
| U | 694 hCoV-19/England/MILK-B3BA6D/2020\|EPI_ISL_679785\|20 | ···················· | ···················· | ···················· | ···················· | ······T············· | ···················· |
| U | 695 hCoV-19/England/MILK-B3BA8B/2020\|EPI_ISL_679786\|20 | ···················· | ···················· | ···················· | ···················· | ···················· | ···················· |
| U | 696 hCoV-19/England/MILK-B3BAD6/2020\|EPI_ISL_679788\|20 | ···················· | ···················· | ···················· | ···················· | ···················· | ···················· |
| U | 697 hCoV-19/England/MILK-B3BBB5/2020\|EPI_ISL_679790\|20 | ···················· | ···················· | ···················· | ···················· | ···················· | ···················· |
| U | 698 hCoV-19/England/MILK-B3BBC4/2020\|EPI_ISL_679791\|20 | ···················· | ···················· | ···················· | ···················· | ···················· | ···················· |
| U | 699 hCoV-19/England/MILK-B3BBD3/2020\|EPI_ISL_679792\|20 | ···················· | ···················· | ···················· | ···················· | ···················· | ···················· |
| U | 700 hCoV-19/England/MILK-B3BBE2/2020\|EPI_ISL_679793\|20 | ···················· | ···················· | ···················· | ···················· | ···················· | ···················· |
| U | 701 hCoV-19/England/MILK-B3BBF1/2020\|EPI_ISL_679794\|20 | ···················· | ···················· | ···················· | ···················· | ···················· | ···················· |
| U | 702 hCoV-19/England/MILK-B3BC0D/2020\|EPI_ISL_679795\|20 | ···················· | ···················· | ···················· | ···················· | ···················· | ···················· |
| U | 703 hCoV-19/England/MILK-B3BC1C/2020\|EPI_ISL_679796\|20 | ···················· | ···················· | ···················· | ···················· | ···················· | ···················· |
| U | 704 hCoV-19/England/MILK-B3BC3A/2020\|EPI_ISL_679797\|20 | ···················· | ···················· | ···················· | ···················· | ······T············· | ···················· |
| U | 705 hCoV-19/England/MILK-B3BC67/2020\|EPI_ISL_679798\|20 | ···················· | ·A·················· | ···················· | ···················· | ···················· | ···A················ |
| U | 706 hCoV-19/England/MILK-B3BC76/2020\|EPI_ISL_679799\|20 | ···················· | ···················· | ···················· | ···················· | ···················· | ···················· |
| U | 707 hCoV-19/Wales/PHWC-48D92A/2020\|EPI_ISL_680700\|2020 | ···················· | ···················· | ···················· | ···················· | ···················· | ···················· |
| U | 708 hCoV-19/Wales/PHWC-48D939/2020\|EPI_ISL_680701\|2020 | ···················· | ···················· | ···················· | ···················· | ···················· | ···················· |
| U | 709 hCoV-19/Wales/PHWC-48D948/2020\|EPI_ISL_680702\|2020 | ···················· | ···················· | ···················· | ···················· | ···················· | ···················· |
| U | 710 hCoV-19/Wales/PHWC-48D975/2020\|EPI_ISL_680704\|2020 | ···················· | ···················· | ···················· | ···················· | ···················· | ···················· |
| U | 712 hCoV-19/Wales/PHWC-48D9B1/2020\|EPI_ISL_680707\|2020 | ···················· | ···················· | ···················· | ···················· | ···················· | ···················· |
| U | 718 hCoV-19/Wales/PHWC-48DA27/2020\|EPI_ISL_680714\|2020 | ···················· | ···················· | ···················· | ···················· | ···················· | ···················· |
| U | 719 hCoV-19/Wales/PHWC-48DA36/2020\|EPI_ISL_680715\|2020 | ···················· | ···················· | ···················· | ···················· | ···················· | ···················· |
| U | 720 hCoV-19/Wales/PHWC-48DA45/2020\|EPI_ISL_680716\|2020 | ···················· | ···················· | ···················· | ···················· | ···················· | ···················· |
| U | 721 hCoV-19/Wales/PHWC-48DA54/2020\|EPI_ISL_680717\|2020 | ···················· | ···················· | ···················· | ···················· | ···················· | ···················· |
| U | 722 hCoV-19/Wales/PHWC-48DA81/2020\|EPI_ISL_680719\|2020 | ···················· | ···················· | ···················· | ···················· | ···················· | ···················· |
| U | 723 hCoV-19/Wales/PHWC-48DA90/2020\|EPI_ISL_680720\|2020 | ···················· | ···················· | ···················· | ···················· | ···················· | ···················· |
| U | 724 hCoV-19/Wales/PHWC-48DAAF/2020\|EPI_ISL_680721\|2020 | ···················· | ···················· | ···················· | ···················· | ···················· | ···················· |
| U | 725 hCoV-19/Wales/PHWC-48DABE/2020\|EPI_ISL_680722\|2020 | ···················· | ···················· | ···················· | ···················· | ···················· | ···················· |
| U | 726 hCoV-19/Wales/PHWC-48DACD/2020\|EPI_ISL_680723\|2020 | ···················· | ···················· | ···················· | ···················· | ···················· | ···················· |
| U | 727 hCoV-19/Wales/PHWC-48DADC/2020\|EPI_ISL_680724\|2020 | ···················· | ···················· | ···················· | ···················· | ···················· | ···················· |
| U | 728 hCoV-19/Wales/PHWC-48DAEB/2020\|EPI_ISL_680725\|2020 | ···················· | ···················· | ···················· | ···················· | ···················· | ···················· |
| U | 729 hCoV-19/Wales/PHWC-48DAFA/2020\|EPI_ISL_680726\|2020 | ···················· | ···················· | ···················· | ···················· | ···················· | ···················· |
| U | 730 hCoV-19/Wales/PHWC-48DB06/2020\|EPI_ISL_680727\|2020 | ···················· | ···················· | ···················· | ···················· | ···················· | ···················· |
| U | 731 hCoV-19/Wales/PHWC-48DB15/2020\|EPI_ISL_680728\|2020 | ···················· | ···················· | ···················· | ···················· | ···················· | ···················· |
| U | 732 hCoV-19/Wales/PHWC-48DB33/2020\|EPI_ISL_680729\|2020 | ···················· | ···················· | ···················· | ···················· | ···················· | ···················· |
| U | 733 hCoV-19/Wales/PHWC-48DB42/2020\|EPI_ISL_680730\|2020 | ···················· | ···················· | ···················· | ···················· | ···················· | ···················· |
| U | 734 hCoV-19/Wales/PHWC-48DB51/2020\|EPI_ISL_680731\|2020 | ···················· | ···················· | ···················· | ···················· | ···················· | ···················· |
| U | 735 hCoV-19/Wales/PHWC-48DB7F/2020\|EPI_ISL_680732\|2020 | ···················· | ···················· | ···················· | ···················· | ···················· | ···················· |
| U | 736 hCoV-19/Wales/PHWC-48DB8E/2020\|EPI_ISL_680733\|2020 | ···················· | ···················· | ···················· | ···················· | ···················· | ···················· |
| U | 737 hCoV-19/Wales/PHWC-48DB9D/2020\|EPI_ISL_680734\|2020 | ···················· | ···················· | ···················· | ···················· | ···················· | ···················· |
| U | 738 hCoV-19/Wales/PHWC-48DBAC/2020\|EPI_ISL_680735\|2020 | ···················· | ···················· | ···················· | ···················· | ···················· | ···················· |
| U | 739 hCoV-19/Wales/PHWC-48DBCA/2020\|EPI_ISL_680737\|2020 | ···················· | ···················· | ···················· | ···················· | ···················· | ···················· |

| | | | | | | | | | |
|---|---|---|---|---|---|---|---|---|---|
| U | 740 | hCoV-19/Wales/PHWC-48DBD9/2020\|EPI_ISL_680738\|2020 | ···················· | ···················· | ····················· | ···················· | ···················· | ···················· | |
| U | 741 | hCoV-19/Wales/PHWC-48DC21/2020\|EPI_ISL_680739\|2020 | ···················· | ···················· | ····················· | ···················· | ···················· | ···················· | |
| U | 742 | hCoV-19/Wales/PHWC-48DE94/2020\|EPI_ISL_680770\|2020 | ···················· | ···················· | ····················· | ···················· | ···················· | ···················· | |
| U | 743 | hCoV-19/Wales/PHWC-48DEA3/2020\|EPI_ISL_680771\|2020 | ···················· | ···················· | ····················· | ···················· | ···················· | ···················· | |
| U | 744 | hCoV-19/Wales/PHWC-48DEB2/2020\|EPI_ISL_680772\|2020 | ···················· | ···················· | ····················· | ···················· | ···················· | N··················· | |
| U | 745 | hCoV-19/Wales/PHWC-48DEC1/2020\|EPI_ISL_680773\|2020 | ···················· | ···················· | ····················· | ···················· | ···················· | ···················· | |
| U | 746 | hCoV-19/Wales/PHWC-48DED0/2020\|EPI_ISL_680774\|2020 | ···················· | ···················· | ····················· | ···················· | ···················· | NNN················· | 1 |
| U | 747 | hCoV-19/Wales/PHWC-48DEEF/2020\|EPI_ISL_680775\|2020 | ···················· | ···················· | ····················· | ···················· | ···················· | ···················· | |
| U | 748 | hCoV-19/Wales/PHWC-48DEFE/2020\|EPI_ISL_680776\|2020 | ···················· | ···················· | ····················· | ···················· | ···················· | ···················· | |
| U | 749 | hCoV-19/Wales/PHWC-48DF0A/2020\|EPI_ISL_680777\|2020 | ···················· | ···················· | ····················· | ···················· | ···················· | ···················· | |
| U | 750 | hCoV-19/Wales/PHWC-48DF19/2020\|EPI_ISL_680778\|2020 | ···················· | ···················· | ····················· | ···················· | ···················· | ···················· | |
| U | 751 | hCoV-19/Wales/PHWC-48DF28/2020\|EPI_ISL_680779\|2020 | ···················· | ···················· | ····················· | ···················· | ···················· | ···················· | |
| U | 752 | hCoV-19/Wales/PHWC-48DF37/2020\|EPI_ISL_680780\|2020 | ···················· | ···················· | ····················· | ···················· | ···················· | ···················· | |
| U | 753 | hCoV-19/Wales/PHWC-48DF46/2020\|EPI_ISL_680781\|2020 | ···················· | ···················· | ····················· | ···················· | ···················· | ···················· | |
| U | 754 | hCoV-19/Wales/PHWC-48DF55/2020\|EPI_ISL_680782\|2020 | ···················· | ···················· | ····················· | ···················· | ···················· | ···················· | |
| U | 755 | hCoV-19/Wales/PHWC-48DF64/2020\|EPI_ISL_680783\|2020 | ···················· | ···················· | ····················· | ···················· | ···················· | ···················· | |
| U | 756 | hCoV-19/Wales/PHWC-48DF73/2020\|EPI_ISL_680784\|2020 | ···················· | ···················· | ····················· | ···················· | ···················· | ···················· | |
| U | 757 | hCoV-19/Wales/PHWC-48DF91/2020\|EPI_ISL_680786\|2020 | ···················· | ···················· | ····················· | ···················· | ···················· | ···················· | |
| U | 758 | hCoV-19/Wales/PHWC-48DFA0/2020\|EPI_ISL_680787\|2020 | ···················· | ···················· | ····················· | ···················· | ···················· | ···················· | |
| U | 759 | hCoV-19/Wales/PHWC-48DFBF/2020\|EPI_ISL_680788\|2020 | ···················· | ···················· | ····················· | ···················· | ···················· | ···················· | |
| U | 760 | hCoV-19/Wales/PHWC-48DFCE/2020\|EPI_ISL_680789\|2020 | ···················· | ···················· | ····················· | ···················· | ···················· | ···················· | |
| U | 761 | hCoV-19/Wales/PHWC-48DFDD/2020\|EPI_ISL_680790\|2020 | ···················· | ···················· | ····················· | ···················· | ···················· | ···················· | |
| U | 762 | hCoV-19/Wales/PHWC-48E006/2020\|EPI_ISL_680793\|2020 | ···················· | ···················· | ····················· | ···················· | ···················· | ···················· | |
| U | 763 | hCoV-19/Wales/PHWC-48E015/2020\|EPI_ISL_680794\|2020 | ···················· | ···················· | ····················· | ···················· | ···················· | ···················· | |
| U | 764 | hCoV-19/Wales/PHWC-48E024/2020\|EPI_ISL_680795\|2020 | ···················· | ···················· | ····················· | ···················· | ···················· | ···················· | |
| U | 765 | hCoV-19/Wales/PHWC-48E033/2020\|EPI_ISL_680796\|2020 | ···················· | ···················· | ····················· | ···················· | ···················· | ···················· | |
| U | 766 | hCoV-19/Wales/PHWC-48E042/2020\|EPI_ISL_680797\|2020 | ···················· | ···················· | ····················· | ···················· | ···················· | ···················· | |
| U | 767 | hCoV-19/Wales/PHWC-48E051/2020\|EPI_ISL_680798\|2020 | ···················· | ···················· | ····················· | ···················· | ···················· | ···················· | |
| U | 768 | hCoV-19/Wales/PHWC-48E060/2020\|EPI_ISL_680799\|2020 | ···················· | ···················· | ····················· | ···················· | ···················· | ···················· | |
| U | 769 | hCoV-19/England/CAMC-B99BD3/2020\|EPI_ISL_673502\|20 | ···················· | ···················· | ····················· | ···················· | ···················· | ···················· | |
| U | 770 | hCoV-19/England/CAMC-B99881/2020\|EPI_ISL_673503\|20 | ···················· | ···················· | ····················· | ···················· | ···················· | ···················· | |
| U | 771 | hCoV-19/England/CAMC-B99D0A/2020\|EPI_ISL_673504\|20 | ···················· | ·A·················· | ····················· | ···················· | ···················· | ···A················ | |
| U | 772 | hCoV-19/England/CAMC-B9A3AD/2020\|EPI_ISL_673505\|20 | ········T··········· | ···················· | ····················· | ···················· | ···················· | ···················· | |
| U | 773 | hCoV-19/England/CAMC-B99DA0/2020\|EPI_ISL_673506\|20 | ···················· | ···················· | ····················· | ···················· | ···················· | ···················· | |
| U | 774 | hCoV-19/England/CAMC-B9C50F/2020\|EPI_ISL_673508\|20 | ···················· | ···················· | ····················· | ···················· | ···················· | ···················· | |
| U | 775 | hCoV-19/England/CAMC-B99FF5/2020\|EPI_ISL_673509\|20 | ···················· | ···················· | ····················· | ···················· | ···················· | ···················· | |
| U | 776 | hCoV-19/England/CAMC-B997B1/2020\|EPI_ISL_673510\|20 | ···················· | ···················· | ····················· | ···················· | ···················· | ···················· | |
| U | 777 | hCoV-19/England/CAMC-B9A176/2020\|EPI_ISL_673511\|20 | ···················· | ···················· | ····················· | ···················· | ···················· | ···················· | |
| U | 778 | hCoV-19/England/CAMC-B99CC1/2020\|EPI_ISL_673512\|20 | ···················· | ·A·················· | ····················· | ···················· | ···················· | ···················· | |
| U | 779 | hCoV-19/England/CAMC-B99872/2020\|EPI_ISL_673514\|20 | ···················· | ···················· | ····················· | ···················· | ···················· | ···················· | |
| U | 780 | hCoV-19/England/CAMC-B99AE5/2020\|EPI_ISL_673515\|20 | ···················· | ···················· | ····················· | ···················· | ···················· | ···················· | |
| U | 781 | hCoV-19/England/CAMC-B99B1F/2020\|EPI_ISL_673516\|20 | ···················· | ···················· | ····················· | ···················· | ···················· | ···················· | |
| U | 782 | hCoV-19/England/CAMC-B9ABE1/2020\|EPI_ISL_673517\|20 | ···················· | ···················· | ····················· | ···················· | ···················· | ···················· | |
| U | 783 | hCoV-19/England/CAMC-B998BE/2020\|EPI_ISL_673519\|20 | ···················· | ···················· | ····················· | ···················· | ···················· | ···················· | |
| U | 784 | hCoV-19/England/CAMC-B99FE6/2020\|EPI_ISL_673520\|20 | ···················· | ···················· | ····················· | ···················· | ···················· | ···················· | |
| U | 785 | hCoV-19/England/CAMC-B9A2A0/2020\|EPI_ISL_673521\|20 | ···················· | ···················· | ····················· | ···················· | ···················· | ···················· | |
| U | 786 | hCoV-19/England/CAMC-B9AA7B/2020\|EPI_ISL_673523\|20 | ···················· | ···················· | ····················· | ···················· | ···················· | ···················· | |
| U | 787 | hCoV-19/England/CAMC-B9972A/2020\|EPI_ISL_673527\|20 | ···················· | ···················· | ····················· | ···················· | ···················· | ···················· | |
| U | 788 | hCoV-19/England/CAMC-B9ABD2/2020\|EPI_ISL_673528\|20 | ···················· | ···················· | ····················· | ···················· | ···················· | ···················· | |
| U | 789 | hCoV-19/England/CAMC-B9A167/2020\|EPI_ISL_673529\|20 | ···················· | ···················· | ····················· | ···················· | ···················· | ···················· | |
| U | 790 | hCoV-19/England/CAMC-B99E34/2020\|EPI_ISL_673532\|20 | ···················· | ···················· | ····················· | ···················· | ······T············· | ···················· | |
| U | 791 | hCoV-19/England/CAMC-B98FE7/2020\|EPI_ISL_673533\|20 | ···················· | ···················· | ····················· | ···················· | ···················· | ···················· | |
| U | 792 | hCoV-19/England/CAMC-B9957B/2020\|EPI_ISL_673535\|20 | ···················· | ···················· | ····················· | ···················· | ···················· | ···················· | |
| U | 793 | hCoV-19/England/CAMC-B9A413/2020\|EPI_ISL_673536\|20 | ···················· | ···················· | ····················· | ···················· | ···················· | ···················· | |
| U | 794 | hCoV-19/England/CAMC-B9A2FB/2020\|EPI_ISL_673537\|20 | ···················· | ···················· | ····················· | ···················· | ···················· | ···················· | |
| U | 795 | hCoV-19/England/CAMC-B9A149/2020\|EPI_ISL_673538\|20 | ···················· | ···················· | ····················· | ···················· | ···················· | ···················· | |
| U | 796 | hCoV-19/England/CAMC-B9A440/2020\|EPI_ISL_673539\|20 | ···················· | ···················· | ····················· | ···················· | ···················· | ···················· | |
| U | 797 | hCoV-19/England/CAMC-B996E1/2020\|EPI_ISL_673541\|20 | ···················· | ···················· | ····················· | ···················· | ···················· | ···················· | |
| U | 798 | hCoV-19/England/CAMC-B99D28/2020\|EPI_ISL_673542\|20 | ···················· | ···················· | ····················· | ···················· | ···················· | ···················· | |
| U | 799 | hCoV-19/England/CAMC-B99B2E/2020\|EPI_ISL_673543\|20 | ···················· | ···················· | ····················· | ···················· | ···················· | ···················· | |
| U | 800 | hCoV-19/England/CAMC-B99353/2020\|EPI_ISL_673544\|20 | ···················· | ···················· | ····················· | ···················· | ···················· | ···················· | |
| U | 801 | hCoV-19/England/CAMC-B99CD0/2020\|EPI_ISL_673545\|20 | ···················· | ···················· | ····················· | ···················· | ···················· | ···················· | |
| U | 802 | hCoV-19/England/CAMC-B99B79/2020\|EPI_ISL_673547\|20 | ···················· | ···················· | ····················· | ···················· | ···················· | ···················· | |
| U | 803 | hCoV-19/England/CAMC-B9956C/2020\|EPI_ISL_673550\|20 | ···················· | ···················· | ····················· | ···················· | ···················· | ···················· | |
| U | 804 | hCoV-19/England/CAMC-B98D92/2020\|EPI_ISL_673551\|20 | ···················· | ···················· | ····················· | ···················· | ···················· | ···················· | |
| U | 805 | hCoV-19/England/CAMC-B9AC1B/2020\|EPI_ISL_673552\|20 | ···················· | ···················· | ····················· | ···················· | ···················· | ···················· | |
| U | 806 | hCoV-19/England/CAMC-B9A000/2020\|EPI_ISL_673554\|20 | ···················· | ···················· | ····················· | ···················· | ···················· | ···················· | |
| U | 807 | hCoV-19/England/CAMC-B99FB9/2020\|EPI_ISL_673555\|20 | ···················· | ···················· | ····················· | ···················· | ···················· | ···················· | |
| U | 808 | hCoV-19/England/CAMC-B995B7/2020\|EPI_ISL_673556\|20 | ········T··········· | ···················· | ····················· | ···················· | ···················· | ···················· | |
| U | 809 | hCoV-19/England/CAMC-B99C58/2020\|EPI_ISL_673557\|20 | ···················· | ···················· | ····················· | ···················· | ···················· | ···················· | |
| U | 810 | hCoV-19/England/CAMC-B98F23/2020\|EPI_ISL_673559\|20 | ···················· | ···················· | ····················· | ···················· | ···················· | ···················· | |
| U | 811 | hCoV-19/England/CAMC-B9A2EC/2020\|EPI_ISL_673561\|20 | ···················· | ···················· | ····················· | ···················· | ···················· | ···················· | |
| U | 812 | hCoV-19/England/CAMC-B992B0/2020\|EPI_ISL_673563\|20 | ···················· | ···················· | ····················· | ···················· | ···················· | ···················· | |
| U | 813 | hCoV-19/England/CAMC-B99BF1/2020\|EPI_ISL_673564\|20 | ···················· | ······A············· | ····················· | ···················· | ···················· | ···················· | |
| U | 814 | hCoV-19/England/CAMC-B99DCE/2020\|EPI_ISL_673565\|20 | ···················· | ···················· | ····················· | ···················· | ···················· | ···················· | |
| U | 815 | hCoV-19/England/CAMC-B9A291/2020\|EPI_ISL_673567\|20 | ···················· | ···················· | ····················· | ···················· | ···················· | ···················· | |
| U | 816 | hCoV-19/England/CAMC-B9A38F/2020\|EPI_ISL_673568\|20 | ···················· | ···················· | ····················· | ···················· | ···················· | ···················· | |
| U | 817 | hCoV-19/England/CAMC-B99EDA/2020\|EPI_ISL_673569\|20 | ···················· | ···················· | ····················· | ···················· | ···················· | ···················· | |
| U | 818 | hCoV-19/England/CAMC-B98DED/2020\|EPI_ISL_673570\|20 | ···················· | ·A·················· | ····················· | ···················· | ···················· | ···················· | |
| U | 819 | hCoV-19/England/CAMC-B9A63B/2020\|EPI_ISL_673571\|20 | ···················· | ···················· | ····················· | ···················· | ···················· | ···················· | |

| | | | | | | | |
|---|---|---|---|---|---|---|---|
| U | 820 hCoV-19/England/CAMC-B9A246/2020\|EPI_ISL_673573\|20 | ···················· | ···················· | ····················· | ···················· | ···················· | ···················· |
| U | 821 hCoV-19/England/CAMC-B99942/2020\|EPI_ISL_673574\|20 | ···················· | ···················· | ····················· | ···················· | ···················· | ···················· |
| U | 822 hCoV-19/England/CAMC-B99EBC/2020\|EPI_ISL_673575\|20 | ···················· | ···················· | ····················· | ···················· | ···················· | ···················· |
| U | 823 hCoV-19/England/CAMC-B98785/2020\|EPI_ISL_673576\|20 | ···················· | ···················· | ····················· | ···················· | ···················· | ···················· |
| U | 824 hCoV-19/England/MILK-B9581C/2020\|EPI_ISL_673577\|20 | ···················· | ···················· | ····················· | ···················· | ···················· | ···················· |
| U | 825 hCoV-19/England/CAMC-B99E8F/2020\|EPI_ISL_673579\|20 | ···················· | ···················· | ····················· | ···················· | ···················· | ···················· |
| U | 826 hCoV-19/England/CAMC-B9A5D4/2020\|EPI_ISL_673580\|20 | ···················· | ···················· | ····················· | ···················· | ···················· | ···················· |
| U | 827 hCoV-19/England/CAMC-B98E17/2020\|EPI_ISL_673581\|20 | ···················· | ···················· | ····················· | ···················· | ···················· | ···················· |
| U | 828 hCoV-19/England/CAMC-B98D1A/2020\|EPI_ISL_673582\|20 | ···················· | ···················· | ····················· | ···················· | ···················· | ···················· |
| U | 829 hCoV-19/England/CAMC-B9A1A3/2020\|EPI_ISL_673583\|20 | ···················· | ···················· | ····················· | ···················· | ···················· | ···················· |
| U | 830 hCoV-19/England/CAMC-B98925/2020\|EPI_ISL_673584\|20 | ···················· | ···················· | ····················· | ···················· | ···················· | ···················· |
| U | 831 hCoV-19/England/CAMC-B99F8C/2020\|EPI_ISL_673585\|20 | ···················· | ···················· | ····················· | ···················· | ···················· | ···················· |
| U | 832 hCoV-19/England/CAMC-B9A5F2/2020\|EPI_ISL_673588\|20 | ···················· | ···················· | ····················· | ···················· | ···················· | ···················· |
| U | 833 hCoV-19/England/CAMC-B9A686/2020\|EPI_ISL_673589\|20 | ···················· | ···················· | ····················· | ···················· | ···················· | ···················· |
| U | 834 hCoV-19/England/CAMC-B98758/2020\|EPI_ISL_673590\|20 | ···················· | ···················· | ····················· | ···················· | ·····T·············· | ···················· |
| U | 835 hCoV-19/England/CAMC-B9A237/2020\|EPI_ISL_673593\|20 | ···················· | ···················· | ····················· | ···················· | ···················· | ···················· |
| U | 836 hCoV-19/England/MILK-B94BD8/2020\|EPI_ISL_673594\|20 | ···················· | ···················· | ····················· | ···················· | ···················· | ···················· |
| U | 837 hCoV-19/England/CAMC-B98C1D/2020\|EPI_ISL_673595\|20 | ···················· | ···················· | ····················· | ···················· | ···················· | ···················· |
| U | 838 hCoV-19/England/CAMC-B9A1EF/2020\|EPI_ISL_673596\|20 | ···················· | ···················· | ····················· | ···················· | ···················· | ···················· |
| U | 839 hCoV-19/England/CAMC-B996F0/2020\|EPI_ISL_673597\|20 | ···················· | ···················· | ····················· | ···················· | ·····T·············· | ···················· |
| U | 840 hCoV-19/England/MILK-B952F0/2020\|EPI_ISL_673599\|20 | ···················· | ···················· | ····················· | ···················· | ···················· | ···················· |
| U | 841 hCoV-19/England/MILK-B8F96D/2020\|EPI_ISL_674800\|20 | ···················· | ···················· | ····················· | ···················· | ···················· | ···················· |
| U | 842 hCoV-19/England/MILK-B8DD54/2020\|EPI_ISL_674803\|20 | ···················· | ···················· | ····················· | ···················· | ···················· | ···················· |
| U | 843 hCoV-19/England/MILK-B8F763/2020\|EPI_ISL_674805\|20 | ···················· | ···················· | ····················· | ···················· | ···················· | ···················· |
| U | 844 hCoV-19/England/QEUH-B7F775/2020\|EPI_ISL_674808\|20 | ···················· | ···················· | ····················· | ···················· | ···················· | ···················· |
| U | 845 hCoV-19/England/QEUH-B7F8DC/2020\|EPI_ISL_674809\|20 | ···················· | ···················· | ····················· | ···················· | ···················· | ···················· |
| U | 846 hCoV-19/England/MILK-B8DFC7/2020\|EPI_ISL_674810\|20 | ···················· | ···················· | ····················· | ···················· | ···················· | ···················· |
| U | 847 hCoV-19/England/MILK-B8E9B9/2020\|EPI_ISL_674811\|20 | ···················· | ···················· | ····················· | ···················· | ···················· | ···················· |
| U | 848 hCoV-19/England/MILK-B8F851/2020\|EPI_ISL_674812\|20 | ···················· | ···················· | ····················· | ···················· | ···················· | ···················· |
| U | 849 hCoV-19/England/MILK-B8E38E/2020\|EPI_ISL_674813\|20 | ···················· | ···················· | ····················· | ···················· | ···················· | ···················· |
| U | 850 hCoV-19/England/MILK-B95955/2020\|EPI_ISL_674814\|20 | ···················· | ···················· | ····················· | ···················· | ···················· | ···················· |
| U | 851 hCoV-19/England/MILK-B976C5/2020\|EPI_ISL_674815\|20 | ···················· | ·A·················· | ····················· | ···················· | ···················· | ···················· |
| U | 852 hCoV-19/England/MILK-B959FB/2020\|EPI_ISL_674816\|20 | ···················· | ···················· | ····················· | ···················· | ···················· | ···················· |
| U | 853 hCoV-19/England/MILK-B9720D/2020\|EPI_ISL_674817\|20 | ···················· | ···················· | ····················· | ···················· | ···················· | ···················· |
| U | 854 hCoV-19/England/MILK-B95EFC/2020\|EPI_ISL_674818\|20 | ···················· | ···················· | ····················· | ···················· | ·····T·············· | ···················· |
| U | 855 hCoV-19/England/MILK-B9762F/2020\|EPI_ISL_674819\|20 | ···················· | ···················· | ····················· | ···················· | ···················· | ···················· |
| U | 856 hCoV-19/England/MILK-B97197/2020\|EPI_ISL_674820\|20 | ···················· | ···················· | ····················· | ···················· | ···················· | ···················· |
| U | 857 hCoV-19/England/MILK-B96D1C/2020\|EPI_ISL_674821\|20 | ···················· | ···················· | ····················· | ···················· | ···················· | ···················· |
| U | 858 hCoV-19/England/MILK-B96B7C/2020\|EPI_ISL_674822\|20 | ···················· | ···················· | ····················· | ···················· | ···················· | ···················· |
| U | 859 hCoV-19/England/MILK-B95C3E/2020\|EPI_ISL_674824\|20 | ···················· | ···················· | ····················· | ···················· | ···················· | ···················· |
| U | 860 hCoV-19/England/MILK-B975AA/2020\|EPI_ISL_674825\|20 | ···················· | ···················· | ····················· | ···················· | ···················· | ···················· |
| U | 861 hCoV-19/England/CAMC-B98239/2020\|EPI_ISL_674826\|20 | ···················· | ···················· | ····················· | ···················· | ···················· | ···················· |
| U | 862 hCoV-19/England/MILK-B96E82/2020\|EPI_ISL_674827\|20 | ···················· | ···················· | ····················· | ···················· | ···················· | ···················· |
| U | 863 hCoV-19/England/CAMC-B981B4/2020\|EPI_ISL_674828\|20 | ···················· | ···················· | ····················· | ···················· | ···················· | ···················· |
| U | 864 hCoV-19/England/MILK-B95AE9/2020\|EPI_ISL_674829\|20 | ···················· | ···················· | ····················· | ···················· | ···················· | ···················· |
| U | 865 hCoV-19/England/MILK-B97416/2020\|EPI_ISL_674830\|20 | ···················· | ···················· | ····················· | ···················· | ···················· | ···················· |
| U | 866 hCoV-19/England/MILK-B95AAD/2020\|EPI_ISL_674831\|20 | ···················· | ·A·················· | ····················· | ···················· | ···················· | ··A················· |
| U | 867 hCoV-19/England/MILK-B9758C/2020\|EPI_ISL_674832\|20 | ···················· | ···················· | ····················· | ···················· | ···················· | ···················· |
| U | 868 hCoV-19/England/MILK-B96DB2/2020\|EPI_ISL_674833\|20 | ···················· | ···················· | ····················· | ···················· | ···················· | ···················· |
| U | 869 hCoV-19/England/CAMC-B97FD9/2020\|EPI_ISL_674834\|20 | ···················· | ···················· | ····················· | ···················· | ···················· | ···················· |
| U | 870 hCoV-19/England/MILK-B972FE/2020\|EPI_ISL_674835\|20 | ···················· | ···················· | ····················· | ···················· | ·····T·············· | ···················· |
| U | 871 hCoV-19/England/MILK-B96E19/2020\|EPI_ISL_674836\|20 | ···················· | ···················· | ····················· | ···················· | ···················· | ···················· |
| U | 872 hCoV-19/England/CAMC-B981F0/2020\|EPI_ISL_674837\|20 | ···················· | ···················· | ····················· | ···················· | ···················· | ···················· |
| U | 873 hCoV-19/England/CAMC-B9849D/2020\|EPI_ISL_674838\|20 | ···················· | ···················· | ····················· | ···················· | ···················· | ···················· |
| U | 874 hCoV-19/England/MILK-B971D3/2020\|EPI_ISL_674839\|20 | ···················· | ···················· | ····················· | ···················· | ···················· | ···················· |
| U | 875 hCoV-19/England/CAMC-B9810F/2020\|EPI_ISL_674840\|20 | ···················· | ···················· | ····················· | ···················· | ···················· | ···················· |
| U | 876 hCoV-19/England/MILK-B95C6B/2020\|EPI_ISL_674841\|20 | ···················· | ···················· | ····················· | ···················· | ···················· | ···················· |
| U | 877 hCoV-19/England/MILK-B971B5/2020\|EPI_ISL_674842\|20 | ···················· | ···················· | ····················· | ···················· | ···················· | ···················· |
| U | 878 hCoV-19/England/MILK-B97346/2020\|EPI_ISL_674844\|20 | ···················· | ···················· | ····················· | ···················· | ···················· | ···················· |
| U | 879 hCoV-19/England/CAMC-B982B1/2020\|EPI_ISL_674845\|20 | ···················· | ···················· | ····················· | ···················· | ···················· | ···················· |
| U | 880 hCoV-19/England/CAMC-B9847F/2020\|EPI_ISL_674846\|20 | ···················· | ·A·················· | ····················· | ···················· | ···················· | ··A················· |
| U | 881 hCoV-19/England/CAMC-B98011/2020\|EPI_ISL_674847\|20 | ···················· | ···················· | ····················· | ···················· | ···················· | ···················· |
| U | 882 hCoV-19/England/MILK-B96D85/2020\|EPI_ISL_674848\|20 | ···················· | ···················· | ····················· | ···················· | ···················· | ···················· |
| U | 883 hCoV-19/England/CAMC-B980A8/2020\|EPI_ISL_674849\|20 | ···················· | ···················· | ····················· | ···················· | ···················· | ···················· |
| U | 884 hCoV-19/England/CAMC-B98424/2020\|EPI_ISL_674850\|20 | ···················· | ·A·················· | ····················· | ···················· | ···················· | ··A················· |
| U | 885 hCoV-19/England/CAMC-B98318/2020\|EPI_ISL_674851\|20 | ···················· | ·A·················· | ····················· | ···················· | ···················· | ··A················· |
| U | 886 hCoV-19/England/MILK-B96E73/2020\|EPI_ISL_674853\|20 | ···················· | ···················· | ····················· | ···················· | ···················· | ···················· |
| U | 887 hCoV-19/England/MILK-B97698/2020\|EPI_ISL_674855\|20 | ···················· | ···················· | ····················· | ···················· | ···················· | ···················· |
| U | 888 hCoV-19/England/MILK-B9622C/2020\|EPI_ISL_674856\|20 | ···················· | ···················· | ····················· | ···················· | ·····T·············· | ···················· |
| U | 889 hCoV-19/England/MILK-B959CE/2020\|EPI_ISL_674857\|20 | ···················· | ···················· | ····················· | ···················· | ···················· | ···················· |
| U | 890 hCoV-19/England/MILK-B9730A/2020\|EPI_ISL_674858\|20 | ···················· | ···················· | ····················· | ···················· | ···················· | ···················· |
| U | 891 hCoV-19/England/CAMC-B9812D/2020\|EPI_ISL_674859\|20 | ···················· | ···················· | ····················· | ···················· | ···················· | ···················· |
| U | 892 hCoV-19/England/MILK-B95CA7/2020\|EPI_ISL_674860\|20 | ···················· | ···················· | ····················· | ···················· | ·····T·············· | ···················· |
| U | 893 hCoV-19/England/MILK-B96DFE/2020\|EPI_ISL_674861\|20 | ···················· | ···················· | ····················· | ···················· | ···················· | ···················· |
| U | 894 hCoV-19/England/CAMC-B9805D/2020\|EPI_ISL_674862\|20 | ···················· | ···················· | ····················· | ···················· | ···················· | ···················· |
| U | 895 hCoV-19/England/MILK-B9767A/2020\|EPI_ISL_674863\|20 | ···················· | ···················· | ····················· | ···················· | ·····T·············· | ···················· |
| U | 896 hCoV-19/England/MILK-B95A16/2020\|EPI_ISL_674864\|20 | ···················· | ·A·················· | ····················· | ···················· | ···················· | ··A················· |
| U | 897 hCoV-19/England/MILK-B96D76/2020\|EPI_ISL_674865\|20 | ···················· | ···················· | ····················· | ···················· | ·····T·············· | ···················· |
| U | 898 hCoV-19/England/CAMC-B97F51/2020\|EPI_ISL_674866\|20 | ···················· | ···················· | ····················· | ···················· | ···················· | ···················· |
| U | 899 hCoV-19/England/CAMC-B98284/2020\|EPI_ISL_674867\|20 | ···················· | ·A·················· | ····················· | ···················· | ···················· | ··A················· |

| | | | | | | | | | |
|---|---|---|---|---|---|---|---|---|---|
| U | 900 | hCoV-19/England/CAMC-B982A2/2020\|EPI_ISL_674868\|20 | ···················· | ···················· | ····················· | ···················· | ···················· | ···················· | |
| U | 901 | hCoV-19/England/MILK-B95946/2020\|EPI_ISL_674869\|20 | ···················· | ···················· | ····················· | ···················· | ···················· | ···················· | |
| U | 902 | hCoV-19/England/MILK-B96DEF/2020\|EPI_ISL_674870\|20 | ···················· | ···················· | ····················· | ···················· | ···················· | ···················· | |
| U | 903 | hCoV-19/England/MILK-B95CE3/2020\|EPI_ISL_674874\|20 | ···················· | ···················· | ····················· | ···················· | ···················· | ···················· | |
| U | 904 | hCoV-19/England/MILK-B97522/2020\|EPI_ISL_674875\|20 | ···················· | ···················· | ····················· | ···················· | ···················· | ···················· | |
| U | 905 | hCoV-19/England/CAMC-B985F4/2020\|EPI_ISL_674876\|20 | ···················· | ···················· | ····················· | ···················· | ······T············· | ···················· | |
| U | 906 | hCoV-19/England/MILK-B970B8/2020\|EPI_ISL_674877\|20 | ···················· | ···················· | ····················· | ···················· | ···················· | ···················· | |
| U | 907 | hCoV-19/England/MILK-B96CA6/2020\|EPI_ISL_674878\|20 | ···················· | ···················· | ····················· | ···················· | ···················· | ···················· | |
| U | 908 | hCoV-19/England/MILK-B977C2/2020\|EPI_ISL_674879\|20 | ···················· | ···················· | ····················· | ···················· | ···················· | ···················· | |
| U | 909 | hCoV-19/England/MILK-B97461/2020\|EPI_ISL_674880\|20 | ···················· | ···················· | ····················· | ···················· | ···················· | ···················· | |
| U | 910 | hCoV-19/England/MILK-B95C98/2020\|EPI_ISL_674881\|20 | ···················· | ···················· | ····················· | ···················· | ···················· | ···················· | |
| U | 911 | hCoV-19/England/CAMC-B97FAC/2020\|EPI_ISL_674883\|20 | ···················· | ···················· | ····················· | ···················· | ···················· | ···················· | |
| U | 912 | hCoV-19/England/MILK-B97434/2020\|EPI_ISL_674884\|20 | ···················· | ···················· | ····················· | ···················· | ···················· | ···················· | |
| U | 913 | hCoV-19/England/MILK-B96EEC/2020\|EPI_ISL_674885\|20 | ···················· | ···················· | ····················· | ···················· | ···················· | ···················· | |
| U | 914 | hCoV-19/England/MILK-B9759B/2020\|EPI_ISL_674886\|20 | ···················· | ···················· | ····················· | ···················· | ···················· | ···················· | |
| U | 915 | hCoV-19/England/MILK-B973FB/2020\|EPI_ISL_674887\|20 | ···················· | ···················· | ····················· | ···················· | ···················· | ···················· | |
| U | 916 | hCoV-19/England/MILK-B96C5B/2020\|EPI_ISL_674889\|20 | ···················· | ···················· | ····················· | ···················· | ···················· | ···················· | |
| U | 917 | hCoV-19/England/MILK-B977E0/2020\|EPI_ISL_674890\|20 | ···················· | ···················· | ····················· | ···················· | ······T············· | ···················· | |
| U | 918 | hCoV-19/England/MILK-B973EC/2020\|EPI_ISL_674892\|20 | ···················· | ···················· | ····················· | ···················· | ···················· | ···················· | |
| U | 919 | hCoV-19/England/CAMC-B98697/2020\|EPI_ISL_674893\|20 | ···················· | ·A·················· | ····················· | ···················· | ···················· | ···A················ | |
| U | 920 | hCoV-19/England/MILK-B970A9/2020\|EPI_ISL_674894\|20 | ···················· | ···················· | ····················· | ···················· | ···················· | ···················· | |
| U | 921 | hCoV-19/England/MILK-B95B13/2020\|EPI_ISL_674895\|20 | ···················· | ···················· | ····················· | ···················· | ···················· | ···················· | |
| U | 922 | hCoV-19/England/CAMC-B98178/2020\|EPI_ISL_674896\|20 | ···················· | ···················· | ····················· | ···················· | ···················· | ···················· | |
| U | 923 | hCoV-19/England/MILK-B9712E/2020\|EPI_ISL_674897\|20 | ···················· | ···················· | ····················· | ···················· | ···················· | ···················· | |
| U | 924 | hCoV-19/England/MILK-B961C5/2020\|EPI_ISL_674898\|20 | ···················· | ···················· | ····················· | ···················· | ······T············· | ···················· | |
| U | 925 | hCoV-19/USA/NY-Wadsworth-200236-01/2020\|EPI_ISL_67 | ···················· | ···················· | ····················· | ···················· | ···················· | ···················· | |
| U | 926 | hCoV-19/USA/NY-Wadsworth-200237-01/2020\|EPI_ISL_67 | ···················· | ···················· | ····················· | ···················· | ···················· | ···················· | |
| U | 927 | hCoV-19/USA/NY-Wadsworth-200238-01/2020\|EPI_ISL_67 | ···················· | ··············C····· | ····················· | ···················· | ···················· | ···················· | |
| U | 928 | hCoV-19/USA/NY-Wadsworth-200239-01/2020\|EPI_ISL_67 | ···················· | ···················· | ····················· | ···················· | ···················· | ···················· | |
| U | 929 | hCoV-19/USA/NY-Wadsworth-200240-01/2020\|EPI_ISL_67 | ···················· | ···················· | ····················· | ···················· | ···················· | ···················· | |
| U | 930 | hCoV-19/USA/NY-Wadsworth-200241-01/2020\|EPI_ISL_67 | ···················· | ···················· | ····················· | ···················· | ···················· | ···················· | |
| U | 931 | hCoV-19/USA/NY-Wadsworth-200243-01/2020\|EPI_ISL_67 | ···················· | ···················· | ····················· | ···················· | ···················· | ···················· | |
| U | 932 | hCoV-19/USA/NY-Wadsworth-201446-01/2020\|EPI_ISL_67 | ···················· | ···················· | ····················· | ···················· | ···················· | ···················· | |
| U | 933 | hCoV-19/USA/NY-Wadsworth-201485-01/2020\|EPI_ISL_67 | ···················· | ···················· | ····················· | ···················· | ···················· | ···················· | |
| U | 934 | hCoV-19/USA/NY-Wadsworth-203545-01/2020\|EPI_ISL_67 | ···················· | ···················· | ····················· | ···················· | ···················· | ···················· | |
| U | 935 | hCoV-19/USA/NY-Wadsworth-203546-01/2020\|EPI_ISL_67 | ···················· | ···················· | ····················· | ···················· | ···················· | ···················· | |
| U | 936 | hCoV-19/USA/NY-Wadsworth-203550-01/2020\|EPI_ISL_67 | ···················· | ···················· | ····················· | ···················· | ···················· | ···················· | |
| U | 937 | hCoV-19/USA/NY-Wadsworth-203562-01/2020\|EPI_ISL_67 | ···················· | ···················· | ····················· | ···················· | ···················· | ···················· | |
| U | 938 | hCoV-19/USA/NY-Wadsworth-204625-01/2020\|EPI_ISL_67 | ···················· | ···················· | ····················· | ···················· | ···················· | ···················· | |
| U | 939 | hCoV-19/USA/NY-Wadsworth-204626-01/2020\|EPI_ISL_67 | ···················· | ···················· | ····················· | ···················· | ···················· | ···················· | |
| U | 940 | hCoV-19/USA/NY-Wadsworth-204627-01/2020\|EPI_ISL_67 | ···················· | ···················· | ····················· | ···················· | ···················· | ···················· | |
| U | 941 | hCoV-19/USA/NY-Wadsworth-204628-01/2020\|EPI_ISL_67 | ···················· | ···················· | ····················· | ···················· | ···················· | ···················· | |
| U | 942 | hCoV-19/USA/NY-Wadsworth-204634-01/2020\|EPI_ISL_67 | ···················· | ···················· | ····················· | ···················· | ···················· | ···················· | |
| U | 943 | hCoV-19/USA/NY-Wadsworth-204636-01/2020\|EPI_ISL_67 | ···················· | ···················· | ····················· | ···················· | ···················· | ···················· | |
| U | 944 | hCoV-19/USA/NY-Wadsworth-207202-01/2020\|EPI_ISL_67 | ···················· | ···················· | ····················· | ···················· | ···················· | ···················· | |
| U | 945 | hCoV-19/USA/NY-Wadsworth-207256-01/2020\|EPI_ISL_67 | ···················· | ···················· | ····················· | ···················· | ···················· | ···················· | |
| U | 946 | hCoV-19/USA/NY-Wadsworth-207860-01/2020\|EPI_ISL_67 | ···················· | ···················· | ····················· | ···················· | ···················· | ···················· | |
| U | 947 | hCoV-19/USA/NY-Wadsworth-209101-01/2020\|EPI_ISL_67 | ···················· | ·A·················· | ····················· | ···················· | ···················· | ···················· | |
| U | 948 | hCoV-19/USA/NY-Wadsworth-210262-01/2020\|EPI_ISL_67 | ···················· | ···················· | ····················· | ···················· | ···················· | ···················· | |
| U | 949 | hCoV-19/USA/NY-Wadsworth-210698-01/2020\|EPI_ISL_67 | ···················· | ···················· | ····················· | ···················· | ···················· | ···················· | |
| U | 950 | hCoV-19/USA/NY-Wadsworth-211722-01/2020\|EPI_ISL_67 | ···················· | ·A·················· | ····················· | ···················· | ···················· | ···················· | |
| U | 951 | hCoV-19/USA/NY-Wadsworth-211749-01/2020\|EPI_ISL_67 | ···················· | ·A·················· | ····················· | ···················· | ···················· | ···················· | |
| U | 952 | hCoV-19/USA/NY-Wadsworth-211750-01/2020\|EPI_ISL_67 | ···················· | ·A·················· | ····················· | ···················· | ···················· | ···················· | |
| U | 953 | hCoV-19/USA/NY-Wadsworth-212770-01/2020\|EPI_ISL_67 | ···················· | ···················· | ····················· | ···················· | ···················· | ···················· | |
| U | 954 | hCoV-19/USA/NY-Wadsworth-215212-01/2020\|EPI_ISL_67 | ···················· | ·A·················· | ····················· | ···················· | ···················· | ···················· | |
| U | 955 | hCoV-19/USA/NY-Wadsworth-224416-01/2020\|EPI_ISL_67 | ···················· | ·A·················· | ····················· | ···················· | ···················· | ···················· | |
| U | 956 | hCoV-19/USA/NY-Wadsworth-224539-01/2020\|EPI_ISL_67 | ···················· | ···················· | ····················· | ···················· | ···················· | ···················· | |
| U | 957 | hCoV-19/USA/NY-Wadsworth-224726-01/2020\|EPI_ISL_67 | ···················· | ···················· | ····················· | ···················· | ···················· | ···················· | |
| U | 958 | hCoV-19/USA/NY-Wadsworth-225049-01/2020\|EPI_ISL_67 | ···················· | ·A·················· | ····················· | ···················· | ···················· | ···················· | |
| U | 959 | hCoV-19/USA/NY-Wadsworth-226476-01/2020\|EPI_ISL_67 | ···················· | ···················· | ····················· | ···················· | ···················· | ···················· | |
| U | 960 | hCoV-19/USA/NY-Wadsworth-226480-01/2020\|EPI_ISL_67 | ···················· | ···················· | ····················· | ···················· | ···················· | ···················· | |
| U | 961 | hCoV-19/USA/NY-Wadsworth-226481-01/2020\|EPI_ISL_67 | ···················· | ···················· | ····················· | ···················· | ···················· | ···················· | |
| U | 962 | hCoV-19/USA/NY-Wadsworth-226482-01/2020\|EPI_ISL_67 | ···················· | ···················· | ····················· | ···················· | ···················· | ···················· | |
| U | 963 | hCoV-19/USA/NY-Wadsworth-226483-01/2020\|EPI_ISL_67 | ···················· | ···················· | ····················· | ···················· | ···················· | ···················· | |
| U | 964 | hCoV-19/USA/NY-Wadsworth-226484-01/2020\|EPI_ISL_67 | ···················· | ·A·················· | ····················· | ···················· | ···················· | ···················· | |
| U | 965 | hCoV-19/USA/NY-Wadsworth-226485-01/2020\|EPI_ISL_67 | ···················· | ···················A | ····················· | ···················· | ···················· | ···················· | 1 |
| U | 966 | hCoV-19/USA/NY-Wadsworth-226486-01/2020\|EPI_ISL_67 | ···················· | ···················· | ····················· | ···················· | ···················· | ···················· | |
| U | 967 | hCoV-19/USA/NY-Wadsworth-226487-01/2020\|EPI_ISL_67 | ···················· | ·A·················· | ····················· | ···················· | ···················· | ···················· | |
| U | 968 | hCoV-19/USA/NY-Wadsworth-226518-01/2020\|EPI_ISL_67 | ···················· | ···················· | ····················· | ···················· | ···················· | ···················· | |
| U | 969 | hCoV-19/Australia/NSW2308/2020\|EPI_ISL_678306\|2020 | ···················· | ···················· | ····················· | ···················· | ···················· | ···················· | |
| U | 970 | hCoV-19/Australia/NSW2380/2020\|EPI_ISL_678307\|2020 | ···················· | ···················· | ····················· | ···················· | ···················· | ···················· | |
| U | 971 | hCoV-19/Australia/NSW2383/2020\|EPI_ISL_678308\|2020 | ···················· | ···················· | ····················· | ···················· | ···················· | ···················· | |
| U | 972 | hCoV-19/Australia/NSW2384/2020\|EPI_ISL_678309\|2020 | ···················· | ···················· | ····················· | ···················· | ···················· | ···················· | |
| U | 973 | hCoV-19/Australia/NSW2387/2020\|EPI_ISL_678310\|2020 | ···················· | ···················· | ····················· | ···················· | ···················· | ···················· | |
| U | 974 | hCoV-19/Australia/NSW2540/2020\|EPI_ISL_678314\|2020 | ···················· | ···················· | ····················· | ···················· | ···················· | ···················· | |
| U | 975 | hCoV-19/Australia/NSW2565/2020\|EPI_ISL_678315\|2020 | ···················· | ···················· | ····················· | ···················· | ···················· | ···················· | |
| U | 976 | hCoV-19/Australia/NSW2567/2020\|EPI_ISL_678316\|2020 | ···················· | ···················· | ····················· | ···················· | ···················· | ···················· | |
| U | 977 | hCoV-19/Belgium/ULG-10898/2020\|EPI_ISL_678440\|2020 | ···················· | ···················· | ····················· | ···················· | ···················· | ···················· | |
| U | 978 | hCoV-19/Belgium/ULG-10900/2020\|EPI_ISL_678442\|2020 | ···················· | ···················· | ····················· | ···················· | ···················· | ···················· | |
| U | 979 | hCoV-19/Belgium/ULG-10901/2020\|EPI_ISL_678443\|2020 | ···················· | ···················· | ····················· | ···················· | ···················· | ···················· | |

| | | | | | | | | |
|---|---|---|---|---|---|---|---|---|
| U | 980 | hCoV-19/Belgium/ULG-10902/2020\|EPI_ISL_678444\|2020 | ···················· | ···················· | ···················· | ···················· | ···················· | ···················· |
| U | 981 | hCoV-19/Belgium/ULG-10903/2020\|EPI_ISL_678445\|2020 | ···················· | ···················· | ···················· | ···················· | ···················· | ···················· |
| U | 982 | hCoV-19/Belgium/ULG-10904/2020\|EPI_ISL_678446\|2020 | ···················· | ···················· | ···················· | ···················· | ···················· | ···················· |
| U | 983 | hCoV-19/Belgium/ULG-10905/2020\|EPI_ISL_678447\|2020 | ···················· | ·A·················· | ···················· | ···················· | ···················· | ···················· |
| U | 984 | hCoV-19/Belgium/ULG-10906/2020\|EPI_ISL_678448\|2020 | ···················· | ···················· | ···················· | ···················· | ···················· | ···················· |
| U | 985 | hCoV-19/Belgium/ULG-10907/2020\|EPI_ISL_678449\|2020 | ···················· | ···················· | ···················· | ···················· | ···················· | ···················· |
| U | 986 | hCoV-19/Belgium/ULG-10917/2020\|EPI_ISL_678458\|2020 | ···················· | ····A··············· | ···················· | ···················· | ···················· | ···················· |
| U | 987 | hCoV-19/England/MILK-B3974A/2020\|EPI_ISL_679637\|20 | ···················· | ···················· | ···················· | ···················· | ···················· | ···················· |
| U | 988 | hCoV-19/England/MILK-B39777/2020\|EPI_ISL_679638\|20 | ···················· | ···················· | ···················· | ···················· | ···················· | ···················· |
| U | 989 | hCoV-19/England/MILK-B39786/2020\|EPI_ISL_679639\|20 | ···················· | ···················· | ···················· | ···················· | ···················· | ···················· |
| U | 990 | hCoV-19/England/MILK-B397C2/2020\|EPI_ISL_679640\|20 | ···················· | ···················· | ···················· | ···················· | ···················· | ···················· |
| U | 991 | hCoV-19/England/MILK-B39829/2020\|EPI_ISL_679642\|20 | ···················· | ···················· | ···················· | ···················· | ···················· | ···················· |
| U | 992 | hCoV-19/England/MILK-B39838/2020\|EPI_ISL_679643\|20 | ···················· | ···················· | ···················· | ···················· | ···················· | ···················· |
| U | 993 | hCoV-19/England/MILK-B39847/2020\|EPI_ISL_679644\|20 | ···················· | ···················· | ···················· | ···················· | ···················· | ···················· |
| U | 994 | hCoV-19/England/MILK-B39856/2020\|EPI_ISL_679645\|20 | ···················· | ···················· | ···················· | ···················· | ···················· | ···················· |
| U | 995 | hCoV-19/England/MILK-B39874/2020\|EPI_ISL_679646\|20 | ···················· | ···················· | ···················· | ···················· | ···················· | ···················· |
| U | 996 | hCoV-19/England/MILK-B39883/2020\|EPI_ISL_679647\|20 | ···················· | ···················· | ···················· | ···················· | ···················· | ···················· |
| U | 997 | hCoV-19/England/MILK-B3B793/2020\|EPI_ISL_679769\|20 | ···················· | ···················· | ···················· | ···················· | ···················· | ···················· |
| U | 998 | hCoV-19/England/MILK-B3B7A2/2020\|EPI_ISL_679770\|20 | ···················· | ···················· | ···················· | ···················· | ···················· | ···················· |
| U | 999 | hCoV-19/England/MILK-B3B7EE/2020\|EPI_ISL_679772\|20 | ···················· | ·A·················· | ···················· | ···················· | ···················· | ···A················ |
| U | 1000 | hCoV-19/England/MILK-B3B7FD/2020\|EPI_ISL_679773\|20 | ···················· | ·A·······A·········· | ···················· | ···················· | ···················· | ···················· |
| U | 1001 | hCoV-19/England/MILK-B3B845/2020\|EPI_ISL_679777\|20 | ···················· | ···················· | ···················· | ···················· | ······T············· | ···················· |
| U | 1002 | hCoV-19/England/MILK-B3B863/2020\|EPI_ISL_679779\|20 | ···················· | ···················· | ···················· | ···················· | ···················· | ···················· |
| U | 1003 | hCoV-19/England/MILK-B3BBA6/2020\|EPI_ISL_679789\|20 | ···················· | ···················· | ···················· | ···················· | ···················· | ···················· |
| U | 1004 | hCoV-19/Wales/PHWC-48D438/2020\|EPI_ISL_680624\|2020 | ···················· | ···················· | ···················· | ···················· | ···················· | ···················· |
| U | 1005 | hCoV-19/Wales/PHWC-48D447/2020\|EPI_ISL_680625\|2020 | ···················· | ···················· | ···················· | ···················· | ···················· | ···················· |
| U | 1006 | hCoV-19/Wales/PHWC-48D483/2020\|EPI_ISL_680629\|2020 | ···················· | ···················· | ···················· | ···················· | ···················· | ···················· |
| U | 1007 | hCoV-19/Wales/PHWC-48DD3D/2020\|EPI_ISL_680750\|2020 | ···················· | ···················· | ···················· | ···················· | ···················· | ···················· |
| U | 1008 | hCoV-19/Wales/PHWC-48DD5B/2020\|EPI_ISL_680752\|2020 | ···················· | ···················· | ···················· | ···················· | ···················· | ···················· |
| U | 1009 | hCoV-19/Wales/PHWC-48DD6A/2020\|EPI_ISL_680753\|2020 | ···················· | ···················· | ···················· | ···················· | ···················· | ···················· |
| U | 1010 | hCoV-19/Wales/PHWC-48DD88/2020\|EPI_ISL_680755\|2020 | ···················· | ···················· | ···················· | ···················· | ···················· | ···················· |
| U | 1012 | hCoV-19/Wales/PHWC-48DDB5/2020\|EPI_ISL_680757\|2020 | ···················· | ···················· | ···················· | ···················· | ···················· | ···················· |
| U | 1013 | hCoV-19/Wales/PHWC-48DDC4/2020\|EPI_ISL_680758\|2020 | ···················· | ···················· | ···················· | ···················· | ···················· | ···················· |
| U | 1014 | hCoV-19/Wales/PHWC-48DDF1/2020\|EPI_ISL_680760\|2020 | ···················· | ···················· | ···················· | ···················· | ···················· | ···················· |
| U | 1015 | hCoV-19/Wales/PHWC-48DE0D/2020\|EPI_ISL_680761\|2020 | ···················· | ···················· | ···················· | ···················· | ···················· | ···················· |
| U | 1016 | hCoV-19/Wales/PHWC-48DE1C/2020\|EPI_ISL_680762\|2020 | ···················· | ···················· | ···················· | ···················· | ···················· | ···················· |
| U | 1017 | hCoV-19/Wales/PHWC-48DE2B/2020\|EPI_ISL_680763\|2020 | ···················· | ···················· | ···················· | ···················· | ···················· | ···················· |
| U | 1018 | hCoV-19/Wales/PHWC-48DE76/2020\|EPI_ISL_680768\|2020 | ···················· | ···················· | ···················· | ···················· | ···················· | ···················· |
| U | 1019 | hCoV-19/Wales/PHWC-48DE85/2020\|EPI_ISL_680769\|2020 | ···················· | ···················· | ···················· | ···················· | ···················· | ···················· |
| U | 1020 | hCoV-19/USA/PA-MGEL-00169/2020\|EPI_ISL_681955\|2020 | ···················· | ···················· | ···················· | ···················· | ···················· | ···················· |
| U | 1021 | hCoV-19/USA/PA-MGEL-00170/2020\|EPI_ISL_681956\|2020 | ···················· | ···················· | ···················· | ···················· | ···················· | ···················· |
| U | 1022 | hCoV-19/USA/PA-MGEL-00173/2020\|EPI_ISL_681957\|2020 | ···················· | ···················· | ···················· | ···················· | ···················· | ···················· |
| U | 1023 | hCoV-19/USA/PA-MGEL-00176/2020\|EPI_ISL_681958\|2020 | ···················· | ···················· | ···················· | ···················· | ···················· | ···················· |
| U | 1024 | hCoV-19/USA/PA-MGEL-00184/2020\|EPI_ISL_681959\|2020 | ···················· | ···················· | ···················· | ···················· | ···················· | ···················· |
| U | 1025 | hCoV-19/USA/NY-Wadsworth-192880-01/2020\|EPI_ISL_67 | ···················· | ···················· | ···················· | ···················· | ···················· | ···················· |
| U | 1026 | hCoV-19/USA/NY-Wadsworth-192882-01/2020\|EPI_ISL_67 | ···················· | ··············C····· | ···················· | ···················· | ···················· | ···················· |
| U | 1027 | hCoV-19/USA/NY-Wadsworth-192883-01/2020\|EPI_ISL_67 | ···················· | ··············C····· | ···················· | ···················· | ···················· | ···················· |
| U | 1028 | hCoV-19/USA/NY-Wadsworth-192884-01/2020\|EPI_ISL_67 | ···················· | ··············C····· | ···················· | ···················· | ···················· | ···················· |
| U | 1029 | hCoV-19/USA/NY-Wadsworth-192885-01/2020\|EPI_ISL_67 | ···················· | ··············C····· | ···················· | ···················· | ···················· | ···················· |
| U | 1030 | hCoV-19/USA/NY-Wadsworth-192886-01/2020\|EPI_ISL_67 | ···················· | ··············C····· | ···················· | ···················· | ···················· | ···················· |
| U | 1031 | hCoV-19/USA/NY-Wadsworth-192887-01/2020\|EPI_ISL_67 | ···················· | ··············C····· | ···················· | ···················· | ···················· | ···················· |
| U | 1032 | hCoV-19/USA/NY-Wadsworth-192891-01/2020\|EPI_ISL_67 | ···················· | ···················· | ···················· | ···················· | ···················· | ···················· |
| U | 1033 | hCoV-19/USA/NY-Wadsworth-192892-01/2020\|EPI_ISL_67 | ···················· | ··············C····· | ···················· | ···················· | ···················· | ···················· |
| U | 1034 | hCoV-19/USA/NY-Wadsworth-192893-01/2020\|EPI_ISL_67 | ···················· | ··············C····· | ···················· | ···················· | ···················· | ···················· |
| U | 1035 | hCoV-19/USA/NY-Wadsworth-192894-01/2020\|EPI_ISL_67 | ···················· | ··············C····· | ···················· | ···················· | ···················· | ···················· |
| U | 1036 | hCoV-19/USA/NY-Wadsworth-192896-01/2020\|EPI_ISL_67 | ···················· | ··············C····· | ···················· | ···················· | ···················· | ···················· |
| U | 1037 | hCoV-19/USA/NY-Wadsworth-192905-01/2020\|EPI_ISL_67 | ···················· | ···················· | ···················· | ···················· | ···················· | ···················· |
| U | 1038 | hCoV-19/USA/NY-Wadsworth-192907-01/2020\|EPI_ISL_67 | ···················· | ···················· | ···················· | ···················· | ···················· | ···················· |
| U | 1039 | hCoV-19/USA/NY-Wadsworth-194530-01/2020\|EPI_ISL_67 | ···················· | ···················· | ···················· | ···················· | ···················· | ···················· |
| U | 1040 | hCoV-19/USA/NY-Wadsworth-194531-01/2020\|EPI_ISL_67 | ···················· | ···················· | ···················· | ···················· | ···················· | ···················· |
| U | 1041 | hCoV-19/USA/NY-Wadsworth-194536-01/2020\|EPI_ISL_67 | ···················· | ···················· | ···················· | ···················· | ···················· | ···················· |
| U | 1042 | hCoV-19/USA/NY-Wadsworth-194538-01/2020\|EPI_ISL_67 | ···················· | ···················· | ···················· | ···················· | ···················· | ···················· |
| U | 1043 | hCoV-19/USA/NY-Wadsworth-194545-01/2020\|EPI_ISL_67 | ···················· | ···················· | ···················· | ···················· | ···················· | ···················· |
| U | 1044 | hCoV-19/USA/NY-Wadsworth-194563-01/2020\|EPI_ISL_67 | ···················· | ···················· | ···················· | ···················· | ···················· | ···················· |
| U | 1045 | hCoV-19/USA/NY-Wadsworth-194568-01/2020\|EPI_ISL_67 | ···················· | ···················· | ···················· | ···················· | ···················· | ···················· |
| U | 1046 | hCoV-19/USA/NY-Wadsworth-194569-01/2020\|EPI_ISL_67 | ···················· | ···················· | ···················· | ···················· | ···················· | ···················· |
| U | 1047 | hCoV-19/USA/NY-Wadsworth-194570-01/2020\|EPI_ISL_67 | ···················· | ···················· | ···················· | ···················· | ···················· | ···················· |
| U | 1048 | hCoV-19/USA/NY-Wadsworth-194579-01/2020\|EPI_ISL_67 | ···················· | ···················· | ···················· | ···················· | ···················· | ···················· |
| U | 1049 | hCoV-19/USA/NY-Wadsworth-194583-01/2020\|EPI_ISL_67 | ···················· | ···················· | ···················· | ···················· | ···················· | ···················· |
| U | 1050 | hCoV-19/USA/NY-Wadsworth-194588-01/2020\|EPI_ISL_67 | ···················· | ···················· | ···················· | ···················· | ···················· | ···················· |
| U | 1051 | hCoV-19/USA/NY-Wadsworth-194589-01/2020\|EPI_ISL_67 | ···················· | ···················· | ···················· | ···················· | ···················· | ···················· |
| U | 1052 | hCoV-19/USA/NY-Wadsworth-194591-01/2020\|EPI_ISL_67 | ···················· | ···················· | ···················· | ···················· | ···················· | ···················· |
| U | 1053 | hCoV-19/USA/NY-Wadsworth-194594-01/2020\|EPI_ISL_67 | ···················· | ···················· | ···················· | ···················· | ···················· | ···················· |
| U | 1054 | hCoV-19/USA/NY-Wadsworth-194595-01/2020\|EPI_ISL_67 | ···················· | ···················· | ···················· | ···················· | ···················· | ···················· |
| U | 1055 | hCoV-19/USA/NY-Wadsworth-199731-01/2020\|EPI_ISL_67 | ···················· | ···················· | ···················· | ···················· | ···················· | ···················· |
| U | 1056 | hCoV-19/USA/NY-Wadsworth-199738-01/2020\|EPI_ISL_67 | ···················· | ···················· | ···················· | ···················· | ···················· | ···················· |
| U | 1057 | hCoV-19/USA/NY-Wadsworth-199741-01/2020\|EPI_ISL_67 | ···················· | ···················· | ···················· | ···················· | ···················· | ···················· |
| U | 1058 | hCoV-19/USA/NY-Wadsworth-199755-01/2020\|EPI_ISL_67 | ···················· | ···················· | ···················· | ···················· | ···················· | ···················· |
| U | 1059 | hCoV-19/USA/NY-Wadsworth-199759-01/2020\|EPI_ISL_67 | ···················· | ···················· | ···················· | ···················· | ···················· | ···················· |
| U | 1060 | hCoV-19/USA/NY-Wadsworth-199763-01/2020\|EPI_ISL_67 | ···················· | ···················· | ···················· | ···················· | ···················· | ···················· |

```
U 1061 hCoV-19/USA/NY-Wadsworth-199767-01/2020|EPI_ISL_67 ···················· ···················· ····················· ···················· ···················· ····················
U 1062 hCoV-19/USA/NY-Wadsworth-200214-01/2020|EPI_ISL_67 ···················· ···················· ····················· ···················· ···················· ····················
U 1063 hCoV-19/USA/NY-Wadsworth-200215-01/2020|EPI_ISL_67 ···················· ···················· ····················· ···················· ···················· ····················
U 1064 hCoV-19/USA/NY-Wadsworth-200216-01/2020|EPI_ISL_67 ···················· ···················· ····················· ···················· ···················· ····················
U 1065 hCoV-19/USA/NY-Wadsworth-200217-01/2020|EPI_ISL_67 ···················· ···················· ····················· ···················· ···················· ····················
U 1066 hCoV-19/USA/NY-Wadsworth-200218-01/2020|EPI_ISL_67 ···················· ···················· ····················· ···················· ···················· ····················
U 1067 hCoV-19/USA/NY-Wadsworth-200219-01/2020|EPI_ISL_67 ···················· ···················· ····················· ···················· ···················· ····················
U 1068 hCoV-19/USA/NY-Wadsworth-200220-01/2020|EPI_ISL_67 ···················· ·R·················· ····················· ···················· ···················· ····················
U 1069 hCoV-19/USA/NY-Wadsworth-200221-01/2020|EPI_ISL_67 ···················· ···················· ····················· ···················· ···················· ····················
U 1070 hCoV-19/USA/NY-Wadsworth-200222-01/2020|EPI_ISL_67 ···················· ···················· ····················· ···················· ···················· ····················
U 1071 hCoV-19/USA/NY-Wadsworth-200223-01/2020|EPI_ISL_67 ···················· ···················· ····················· ···················· ···················· ····················
U 1072 hCoV-19/USA/NY-Wadsworth-200225-01/2020|EPI_ISL_67 ···················· ···················· ····················· ···················· ···················· ····················
U 1073 hCoV-19/USA/NY-Wadsworth-200226-01/2020|EPI_ISL_67 ···················· ··············C····· ····················· ···················· ···················· ····················
U 1074 hCoV-19/USA/NY-Wadsworth-200227-01/2020|EPI_ISL_67 ···················· ···················· ····················· ···················· ···················· ····················
U 1075 hCoV-19/USA/NY-Wadsworth-200228-01/2020|EPI_ISL_67 ···················· ···················· ····················· ···················· ···················· ····················
U 1076 hCoV-19/USA/NY-Wadsworth-200229-01/2020|EPI_ISL_67 ···················· ··············C····· ····················· ···················· ···················· ····················
U 1077 hCoV-19/USA/NY-Wadsworth-200230-01/2020|EPI_ISL_67 ···················· ···················· ····················· ···················· ···················· ····················
U 1078 hCoV-19/USA/NY-Wadsworth-200231-01/2020|EPI_ISL_67 ···················· ···················· ····················· ···················· ···················· ····················
U 1079 hCoV-19/USA/NY-Wadsworth-200232-01/2020|EPI_ISL_67 ···················· ··············C····· ····················· ···················· ···················· ····················
U 1080 hCoV-19/USA/NY-Wadsworth-200233-01/2020|EPI_ISL_67 ···················· ··············C····· ····················· ···················· ···················· ····················
U 1081 hCoV-19/USA/NY-Wadsworth-200234-01/2020|EPI_ISL_67 ···················· ···················· ····················· ···················· ···················· ····················
U 1082 hCoV-19/USA/NY-Wadsworth-200235-01/2020|EPI_ISL_67 ···················· ···················· ····················· ···················· ···················· ····················
U 1083 hCoV-19/Australia/NSW2735/2020|EPI_ISL_678340|2020 ···················· ···················· ····················· ···················· ···················· ····················
U 1084 hCoV-19/Australia/NSW2743/2020|EPI_ISL_678341|2020 ···················· ···················· ····················· ···················· ···················· ····················
U 1085 hCoV-19/Australia/NSW2747/2020|EPI_ISL_678342|2020 ···················· ···················· ····················· ···················· ···················· ····················
U 1086 hCoV-19/Australia/NSW2760/2020|EPI_ISL_678343|2020 ···················· ···················· ····················· ···················· ···················· ····················
U 1087 hCoV-19/Australia/NSW2776/2020|EPI_ISL_678345|2020 ···················· ···················· ····················· ···················· ···················· ····················
U 1088 hCoV-19/Australia/NSW2821/2020|EPI_ISL_678351|2020 ···················· ···················· ····················· ···················· ···················· ····················
U 1089 hCoV-19/Australia/NSW2831/2020|EPI_ISL_678354|2020 ···················· ···················· ····················· ···················· ···················· ····················
U 1090 hCoV-19/Australia/NSW2871/2020|EPI_ISL_678358|2020 ···················· ·A·················· ····················· ···················· ···················· ···A················
U 1091 hCoV-19/Australia/NSW2885/2020|EPI_ISL_678359|2020 ···················· ···················· ····················· ···················· ···················· ····················
U 1092 hCoV-19/Australia/NSW2887/2020|EPI_ISL_678360|2020 ···················· ···················· ····················· ···················· ···················· ····················
U 1093 hCoV-19/Australia/NSW2894/2020|EPI_ISL_678361|2020 ···················· ···················· ····················· ···················· ···················· ····················
U 1094 hCoV-19/Australia/NSW2899/2020|EPI_ISL_678363|2020 ···················· ···················· ····················· ···················· ···················· ····················
U 1095 hCoV-19/Australia/NSW2907/2020|EPI_ISL_678365|2020 ···················· ·A·················· ····················· ···················· ···················· ····················
U 1096 hCoV-19/Australia/NSW2910/2020|EPI_ISL_678366|2020 ···················· ·A·················· ····················· ···················· ···················· ···A················
U 1097 hCoV-19/Australia/NSW2914/2020|EPI_ISL_678367|2020 ···················· ···················· ····················· ···················· ···················· ····················
U 1098 hCoV-19/Australia/NSW2915/2020|EPI_ISL_678368|2020 ···················· ···················· ····················· ···················· ···················· ····················
U 1099 hCoV-19/Australia/NSW2938/2020|EPI_ISL_678370|2020 ···················· ···················· ····················· ···················· ···················· ····················
U 1100 hCoV-19/Australia/NSW2939/2020|EPI_ISL_678371|2020 ···················· ···················· ····················· ···················· ···················· ····················
U 1101 hCoV-19/Australia/NSW2940/2020|EPI_ISL_678372|2020 ···················· ···················· ····················· ···················· ···················· ····················
U 1102 hCoV-19/Australia/NSW2941/2020|EPI_ISL_678373|2020 ···················· ···················· ····················· ···················· ···················· ····················
U 1103 hCoV-19/Australia/NSW2942/2020|EPI_ISL_678374|2020 ···················· ···················· ····················· ···················· ···················· ····················
U 1104 hCoV-19/Australia/NSW2943/2020|EPI_ISL_678375|2020 ···················· ···················· ····················· ···················· ···················· ····················
U 1105 hCoV-19/Australia/NSW2944/2020|EPI_ISL_678376|2020 ···················· ···················· ····················· ···················· ···················· ····················
U 1106 hCoV-19/Australia/NSW2947/2020|EPI_ISL_678377|2020 ···················· ···················· ····················· ···················· ···················· ····················
U 1107 hCoV-19/Australia/NSW2949/2020|EPI_ISL_678378|2020 ···················· ···················· ····················· ···················· ···················· ····················
U 1108 hCoV-19/Australia/NSW2953/2020|EPI_ISL_678379|2020 ···················· ···················· ····················· ···················· ···················· ····················
U 1109 hCoV-19/Australia/NSW2954/2020|EPI_ISL_678380|2020 ···················· ·A·················· ····················· ···················· ···················· ···A················
U 1110 hCoV-19/Australia/NSW2955/2020|EPI_ISL_678381|2020 ···················· ···················· ····················· ···················· ···················· ····················
U 1111 hCoV-19/Australia/NSW3075/2020|EPI_ISL_678383|2020 ···················· ···················· ····················· ···················· ···················· ····················
U 1112 hCoV-19/Hungary/UD-94339/2020|EPI_ISL_678387|2020- ···················· ···················· ····················· ···················· ···················· ····················
U 1113 hCoV-19/Hungary/UD-94346/2020|EPI_ISL_678388|2020- ···················· ···················· ····················· ···················· ···················· ····················
U 1114 hCoV-19/Hungary/UD-94347/2020|EPI_ISL_678389|2020- ···················· ···················· ····················· ···················· ···················· ····················
U 1115 hCoV-19/Hungary/UD-94349/2020|EPI_ISL_678390|2020- ···················· ···················· ····················· ···················· ···················· ····················
U 1116 hCoV-19/Hungary/UD-94353/2020|EPI_ISL_678391|2020- ···················· ···················· ····················· ···················· ···················· ····················
U 1117 hCoV-19/Hungary/UD-94354/2020|EPI_ISL_678392|2020- ···················· ···················· ····················· ···················· ···················· ····················
U 1118 hCoV-19/Hungary/UD-94357/2020|EPI_ISL_678393|2020- ···················· ···················· ····················· ···················· ···················· ····················
U 1119 hCoV-19/Hungary/UD-94381/2020|EPI_ISL_678394|2020- ···················· ···················· ····················· ···················· ···················· ····················
U 1120 hCoV-19/Hungary/UD-94387/2020|EPI_ISL_678395|2020- ···················· ···················· ····················· ···················· ···················· ····················
U 1121 hCoV-19/Hungary/UD-94400/2020|EPI_ISL_678396|2020- ···················· ···················· ····················· ···················· ···················· ····················
U 1122 hCoV-19/Romania/ROSV-10281/2020|EPI_ISL_678397|202 ···················· ···················· ····················· ···················· ···················· ····················
U 1123 hCoV-19/Romania/ROSV-1815/2020|EPI_ISL_678398|2020 ···················· ···················· ····················· ···················· ···················· ····················
U 1124 hCoV-19/Belgium/ULG-10855/2020|EPI_ISL_678399|2020 ···················· ···················· ····················· ···················· ···················· ····················
U 1125 hCoV-19/England/LOND-DC595/2020|EPI_ISL_679623|202 ···················· ···················· ····················· ···················· ···················· ····················
U 1126 hCoV-19/England/LOND-DC5A4/2020|EPI_ISL_679624|202 ···················· ···················· ····················· ···················· ······T············· ····················
U 1127 hCoV-19/England/LOND-DC5B3/2020|EPI_ISL_679625|202 ···················· ···················· ····················· ···················· ···················· ····················
U 1128 hCoV-19/England/MILK-A1927D/2020|EPI_ISL_679627|20 ···················· ···················· ····················· ···················· ···················· ····················
U 1129 hCoV-19/England/MILK-B39610/2020|EPI_ISL_679629|20 ···················· ·A·················· ····················· ···················· ···················· ····················
U 1130 hCoV-19/England/MILK-B3962F/2020|EPI_ISL_679630|20 ···················· ···················· ····················· ···················· ······T············· ····················
U 1131 hCoV-19/England/MILK-B3963E/2020|EPI_ISL_679631|20 ···················· ···················· ····················· ···················· ···················· ····················
U 1132 hCoV-19/England/MILK-B3966B/2020|EPI_ISL_679632|20 ···················· ·A·················· ····················· ···················· ···················· ····················
U 1133 hCoV-19/England/MILK-B39689/2020|EPI_ISL_679633|20 ···················· ···················· ····················· ···················· ···················· ····················
U 1134 hCoV-19/England/MILK-B396C5/2020|EPI_ISL_679634|20 ···················· ···················· ····················· ···················· ···················· ····················
U 1135 hCoV-19/England/MILK-B396D4/2020|EPI_ISL_679635|20 ···················· ···················· ····················· ···················· ···················· ····················
U 1136 hCoV-19/England/MILK-B3970E/2020|EPI_ISL_679636|20 ···················· ···················· ····················· ···················· ···················· ····················
U 1137 hCoV-19/England/MILK-B39E27/2020|EPI_ISL_679670|20 ···················· ···················· ····················· ···················· ···················· ····················
U 1138 hCoV-19/England/MILK-B39E36/2020|EPI_ISL_679671|20 ···················· ···················· ····················· ···················· ···················· ····················
U 1139 hCoV-19/England/MILK-B39E45/2020|EPI_ISL_679672|20 ···················· ···················· ····················· ···················· ···················· ····················
U 1140 hCoV-19/England/MILK-B39E63/2020|EPI_ISL_679673|20 ···················· ·A·······A·········· ····················· ···················· ···················· ····················
```

| | | | | | | | | | |
|---|---|---|---|---|---|---|---|---|---|
| U | 1141 | hCoV-19/England/MILK-B39E81/2020\|EPI_ISL_679674\|20 | ···················· | ···················· | ···················· | ···················· | ···················· | ···················· | |
| U | 1142 | hCoV-19/England/MILK-B39E90/2020\|EPI_ISL_679675\|20 | ···················· | ···················· | ···················· | ···················· | ···················· | ···················· | |
| U | 1143 | hCoV-19/England/MILK-B3A099/2020\|EPI_ISL_679676\|20 | ···················· | ···················· | ···················· | ···················· | ···················· | ···················· | |
| U | 1144 | hCoV-19/England/MILK-B3A0A8/2020\|EPI_ISL_679677\|20 | ···················· | ···················· | ···················· | ···················· | ···················· | ···················· | |
| U | 1145 | hCoV-19/England/MILK-B3A0B7/2020\|EPI_ISL_679678\|20 | ···················· | ···················· | ···················· | ···················· | ···················· | ···················· | |
| U | 1146 | hCoV-19/England/MILK-B3A0D5/2020\|EPI_ISL_679680\|20 | ···················· | ···················· | ···················· | ···················· | ···················· | ···A················ | |
| U | 1147 | hCoV-19/England/MILK-B3A0F3/2020\|EPI_ISL_679681\|20 | ···················· | ···················· | ···················· | ···················· | ···················· | ···················· | |
| U | 1148 | hCoV-19/England/MILK-B3A10F/2020\|EPI_ISL_679682\|20 | ···················· | ···················· | ···················· | ···················· | ······T············· | ···················· | |
| U | 1149 | hCoV-19/England/MILK-B3A12D/2020\|EPI_ISL_679683\|20 | ···················· | ···················· | ···················· | ···················· | ···················· | ···A················ | |
| U | 1150 | hCoV-19/England/MILK-B3A13C/2020\|EPI_ISL_679684\|20 | ···················· | ···················· | ···················· | ···················· | ···················· | ···················· | |
| U | 1151 | hCoV-19/England/MILK-B3A14B/2020\|EPI_ISL_679685\|20 | ···················· | ···················· | ···················· | ···················· | ········A··········· | ···················· | |
| U | 1152 | hCoV-19/England/MILK-B3A846/2020\|EPI_ISL_679686\|20 | ···················· | ···················· | ···················· | ···················· | ···················· | ···················· | |
| U | 1153 | hCoV-19/England/MILK-B3A864/2020\|EPI_ISL_679687\|20 | ···················· | ···················· | ···················· | ···················· | ···················· | ···················· | |
| U | 1154 | hCoV-19/England/MILK-B3A907/2020\|EPI_ISL_679689\|20 | ···················· | ···················· | ···················· | ···················· | ···················· | ···················· | |
| U | 1155 | hCoV-19/England/MILK-B3A916/2020\|EPI_ISL_679690\|20 | ···················· | ···················· | ···················· | ···················· | ···················· | ···················· | |
| U | 1156 | hCoV-19/England/MILK-B3A925/2020\|EPI_ISL_679691\|20 | ···················· | ···················· | ···················· | ···················· | ···················· | ···················· | |
| U | 1157 | hCoV-19/England/MILK-B3A934/2020\|EPI_ISL_679692\|20 | ···················· | ···················· | ···················· | ···················· | ······T············· | ···················· | |
| U | 1158 | hCoV-19/England/MILK-B3A961/2020\|EPI_ISL_679693\|20 | ···················· | ···················· | ···················· | ···················· | ···················· | ···················· | |
| U | 1159 | hCoV-19/England/MILK-B3A98F/2020\|EPI_ISL_679694\|20 | ···················· | ···················· | ···················· | ···················· | ······T············· | ···············A···· | |
| U | 1160 | hCoV-19/England/MILK-B3A99E/2020\|EPI_ISL_679695\|20 | ···················· | ···················· | ···················· | ···················· | ···················· | ···················· | |
| U | 1161 | hCoV-19/England/MILK-B3A9AD/2020\|EPI_ISL_679696\|20 | ···················· | ···················· | ···················· | ···················· | ···················· | ···················· | |
| U | 1162 | hCoV-19/England/MILK-B3A9CB/2020\|EPI_ISL_679698\|20 | ···················· | ···················· | ···················· | ···················· | ···················· | ···················· | |
| U | 1163 | hCoV-19/England/MILK-B3A9DA/2020\|EPI_ISL_679699\|20 | ···················· | ···················· | ···················· | ···················· | ···················· | ···················· | |
| U | 1164 | hCoV-19/Wales/PHWC-48D2A7/2020\|EPI_ISL_680600\|2020 | ···················· | ···················· | ···················· | ···················· | ···················· | ···················· | |
| U | 1165 | hCoV-19/Wales/PHWC-48D2B6/2020\|EPI_ISL_680601\|2020 | ···················· | ···················· | ···················· | ···················· | ···················· | ···················· | |
| U | 1166 | hCoV-19/Wales/PHWC-48D2D4/2020\|EPI_ISL_680603\|2020 | ···················· | ···················· | ···················· | ···················· | ···················· | ···················· | |
| U | 1167 | hCoV-19/Wales/PHWC-48D2E3/2020\|EPI_ISL_680604\|2020 | ···················· | ···················· | ···················· | ···················· | ···················· | ···················· | |
| U | 1168 | hCoV-19/Wales/PHWC-48D2F2/2020\|EPI_ISL_680605\|2020 | ···················· | ···················· | ···················· | ···················· | ···················· | ···················· | |
| U | 1169 | hCoV-19/Wales/PHWC-48D30E/2020\|EPI_ISL_680606\|2020 | ···················· | ···················· | ···················· | ···················· | ···················· | ···················· | |
| U | 1170 | hCoV-19/Wales/PHWC-48D31D/2020\|EPI_ISL_680607\|2020 | ···················· | ···················· | ···················· | ···················· | ···················· | ···················· | |
| U | 1171 | hCoV-19/Wales/PHWC-48D32C/2020\|EPI_ISL_680608\|2020 | ···················· | ···················· | ···················· | ···················· | ···················· | ···················· | |
| U | 1172 | hCoV-19/Wales/PHWC-48D33B/2020\|EPI_ISL_680609\|2020 | ···················· | ···················· | ···················· | ···················· | ···················· | ···················· | |
| U | 1173 | hCoV-19/Wales/PHWC-48D34A/2020\|EPI_ISL_680610\|2020 | ···················· | ···················· | ···················· | ···················· | ···················· | ···················· | |
| U | 1174 | hCoV-19/Wales/PHWC-48D368/2020\|EPI_ISL_680612\|2020 | ···················· | ···················· | ···················· | ···················· | ···················· | ···················· | |
| U | 1175 | hCoV-19/Wales/PHWC-48D377/2020\|EPI_ISL_680613\|2020 | ···················· | ···················· | ···················· | ···················· | ···················· | ···················· | |
| U | 1176 | hCoV-19/Wales/PHWC-48D386/2020\|EPI_ISL_680614\|2020 | ···················· | ···················· | ···················· | ···················· | ···················· | ···················· | |
| U | 1177 | hCoV-19/Wales/PHWC-48D395/2020\|EPI_ISL_680615\|2020 | ···················· | ···················· | ···················· | ···················· | ···················· | ···················· | |
| U | 1178 | hCoV-19/Wales/PHWC-48D3A4/2020\|EPI_ISL_680616\|2020 | ···················· | ···················· | ···················· | ···················· | ···················· | ···················· | |
| U | 1179 | hCoV-19/Wales/PHWC-48D3B3/2020\|EPI_ISL_680617\|2020 | ···················· | ···················· | ···················· | ···················· | ···················· | ···················· | |
| U | 1180 | hCoV-19/Wales/PHWC-48D3C2/2020\|EPI_ISL_680618\|2020 | ···················· | ···················· | ···················· | ···················· | ···················· | ···················· | |
| U | 1181 | hCoV-19/Wales/PHWC-48D3E0/2020\|EPI_ISL_680620\|2020 | ···················· | ···················· | ···················· | ···················· | ···················· | ···················· | |
| U | 1182 | hCoV-19/Wales/PHWC-48D3FF/2020\|EPI_ISL_680621\|2020 | ···················· | ···················· | ···················· | ···················· | ···················· | ···················· | |
| U | 1183 | hCoV-19/Wales/PHWC-48D41A/2020\|EPI_ISL_680622\|2020 | ···················· | ···················· | ···················· | ···················· | ···················· | ···················· | |
| U | 1184 | hCoV-19/Wales/PHWC-48D429/2020\|EPI_ISL_680623\|2020 | ···················· | ···················· | ···················· | ···················· | ···················· | ···················· | |
| U | 1185 | hCoV-19/Wales/PHWC-48D474/2020\|EPI_ISL_680628\|2020 | ···················· | ···················· | ···················· | ···················· | ···················· | ···················· | |
| U | 1186 | hCoV-19/Wales/PHWC-48D68D/2020\|EPI_ISL_680660\|2020 | ···················· | ···················· | ···················· | ···················· | ···················· | ···················· | |
| U | 1187 | hCoV-19/Wales/PHWC-48D6BA/2020\|EPI_ISL_680662\|2020 | ···················· | ···················· | ···················· | ···················· | ···················· | ···················· | |
| U | 1188 | hCoV-19/Wales/PHWC-48D6D8/2020\|EPI_ISL_680664\|2020 | ···················· | ···················· | ···················· | ···················· | ···················· | ···················· | |
| U | 1189 | hCoV-19/Wales/PHWC-48D6E7/2020\|EPI_ISL_680665\|2020 | ···················· | ···················· | ···················· | ···················· | ···················· | ···················· | |
| U | 1190 | hCoV-19/Wales/PHWC-48D6F6/2020\|EPI_ISL_680666\|2020 | ···················· | ···················· | ···················· | ···················· | ···················· | ···················· | |
| U | 1191 | hCoV-19/Wales/PHWC-48D720/2020\|EPI_ISL_680669\|2020 | ···················· | ···················· | ···················· | ···················· | ···················· | ···················· | |
| U | 1192 | hCoV-19/Wales/PHWC-48D73F/2020\|EPI_ISL_680670\|2020 | ···················· | ···················· | ···················· | ···················· | ···················· | ···················· | |
| U | 1193 | hCoV-19/Wales/PHWC-48D75D/2020\|EPI_ISL_680672\|2020 | ···················· | ···················· | ···················· | ···················· | ···················· | ···················· | |
| U | 1194 | hCoV-19/Wales/PHWC-48D78A/2020\|EPI_ISL_680675\|2020 | ···················· | ···················· | ···················· | ···················· | ···················· | ···················· | |
| U | 1195 | hCoV-19/Wales/PHWC-48D7A8/2020\|EPI_ISL_680677\|2020 | ···················· | ···················· | ···················· | ···················· | ···················· | ···················· | |
| U | 1196 | hCoV-19/Wales/PHWC-48D7B7/2020\|EPI_ISL_680678\|2020 | ···················· | ···················· | ···················· | ···················· | ···················· | ···················· | |
| U | 1197 | hCoV-19/Wales/PHWC-48D7C6/2020\|EPI_ISL_680679\|2020 | ···················· | ···················· | ···················· | ···················· | ···················· | ···················· | |
| U | 1198 | hCoV-19/Wales/PHWC-48D7D5/2020\|EPI_ISL_680680\|2020 | ···················· | ···················· | ···················· | ···················· | ···················· | ···················· | |
| U | 1199 | hCoV-19/Wales/PHWC-48D7E4/2020\|EPI_ISL_680681\|2020 | ···················· | ···················· | ···················· | ···················· | ···················· | ···················· | |
| U | 1201 | hCoV-19/Wales/PHWC-48D80F/2020\|EPI_ISL_680683\|2020 | ···················· | ···················· | ···················· | ···················· | ···················· | ···················· | |
| U | 1202 | hCoV-19/Wales/PHWC-48D81E/2020\|EPI_ISL_680684\|2020 | ···················· | ···················· | ···················· | ···················· | ···················· | ···················· | |
| U | 1203 | hCoV-19/Wales/PHWC-48D82D/2020\|EPI_ISL_680685\|2020 | ···················· | ···················· | ···················· | ···················· | ···················· | ···················· | |
| U | 1204 | hCoV-19/Wales/PHWC-48D84B/2020\|EPI_ISL_680687\|2020 | ···················· | ···················· | ···················· | ···················· | ···················· | ···················· | |
| U | 1205 | hCoV-19/Wales/PHWC-48D85A/2020\|EPI_ISL_680688\|2020 | ···················· | ···················· | ···················· | ···················· | ···················· | ···················· | |
| U | 1206 | hCoV-19/Wales/PHWC-48D869/2020\|EPI_ISL_680689\|2020 | ···················· | ···················· | ···················· | ···················· | ···················· | ···················· | |
| U | 1207 | hCoV-19/Wales/PHWC-48D878/2020\|EPI_ISL_680690\|2020 | ···················· | ···················· | ···················· | ···················· | ···················· | ···················· | |
| U | 1208 | hCoV-19/Wales/PHWC-48D887/2020\|EPI_ISL_680691\|2020 | ···················· | ···················· | ···················· | ···················· | ···················· | ···················· | |
| U | 1209 | hCoV-19/Wales/PHWC-48D896/2020\|EPI_ISL_680692\|2020 | ···················· | ···················· | ···················· | ···················· | ···················· | ···················· | |
| U | 1210 | hCoV-19/Wales/PHWC-48D8A5/2020\|EPI_ISL_680693\|2020 | ···················· | ···················· | ···················· | ···················· | ···················· | ···················· | |
| U | 1211 | hCoV-19/Wales/PHWC-48D8B4/2020\|EPI_ISL_680694\|2020 | ···················· | ···················· | ···················· | ···················· | ···················· | ···················· | |
| U | 1212 | hCoV-19/Wales/PHWC-48D8C3/2020\|EPI_ISL_680695\|2020 | ···················· | ···················· | ···················· | ···················· | ···················· | ···················· | |
| U | 1213 | hCoV-19/Wales/PHWC-48D8D2/2020\|EPI_ISL_680696\|2020 | ···················· | ···················· | ···················· | ···················· | ···················· | ···················· | |
| U | 1214 | hCoV-19/Wales/PHWC-48D8E1/2020\|EPI_ISL_680697\|2020 | ···················· | ···················· | ···················· | ···················· | ···················· | ···················· | |
| U | 1215 | hCoV-19/Wales/PHWC-48D8F0/2020\|EPI_ISL_680698\|2020 | ···················· | ···················· | ···················· | ···················· | ···················· | ···················· | |
| U | 1216 | hCoV-19/Wales/PHWC-48D90C/2020\|EPI_ISL_680699\|2020 | ···················· | ···················· | ···················· | ···················· | ···················· | ···················· | |
| U | 1217 | hCoV-19/Ireland/D-NVRL-20Z06674/2020\|EPI_ISL_68190 | ···················· | ···················A | ···················· | ···················· | ···················· | ···················· | 1 |
| U | 1218 | hCoV-19/Ireland/WW-NVRL-20Z06427/2020\|EPI_ISL_6819 | ···················· | ···················· | ···················· | ···················· | ···················· | ···················· | |
| U | 1219 | hCoV-19/Ireland/MO-NVRL-20Z04473/2020\|EPI_ISL_6819 | ···················· | ···················· | ···················· | ···················· | ···················· | ···················· | |
| U | 1220 | hCoV-19/Ireland/D-NVRL-20Z06800/2020\|EPI_ISL_68190 | ···················· | ···················· | ···················· | ···················· | ···················· | ···················· | |
| U | 1221 | hCoV-19/Ireland/LH-NVRL-20Z06935/2020\|EPI_ISL_6819 | ···················· | ···················· | ···················· | ···················· | ···················· | ···················· | |

```
U   1222 hCoV-19/Ireland/D-NVRL-20Z06235/2020|EPI_ISL_68190   ····················  ····················  ·····················  ····················  ····················  ····················
U   1223 hCoV-19/Ireland/CO-NVRL-20G49893/2020|EPI_ISL_6819   ····················  ····················  ·····················  ····················  ····················  ····················
U   1224 hCoV-19/Ireland/MH-NVRL-20G49920/2020|EPI_ISL_6819   ····················  ····················  ·····················  ····················  ····················  ····················
U   1225 hCoV-19/Ireland/CO-NVRL-20G45661/2020|EPI_ISL_6819   ····················  ····················  ·····················  ····················  ····················  ····················
U   1226 hCoV-19/Ireland/MO-NVRL-20G45847/2020|EPI_ISL_6819   ····················  ····················  ·····················  ····················  ····················  ····················
U   1227 hCoV-19/Ireland/KK-NVRL-20G46944/2020|EPI_ISL_6819   ····················  ····················  ·····················  ····················  ····················  ····················
U   1228 hCoV-19/Ireland/MH-NVRL-20G46910/2020|EPI_ISL_6819   ····················  ····················  ·····················  ····················  ····················  ····················
U   1229 hCoV-19/Ireland/MH-NVRL-20G43888/2020|EPI_ISL_6819   ····················  ····················  ·····················  ····················  ····················  ····················
U   1230 hCoV-19/Ireland/KK-NVRL-20G43871/2020|EPI_ISL_6819   ····················  ····················  ·····················  ····················  ····················  ····················
U   1231 hCoV-19/Ireland/D-NVRL-20G47385/2020|EPI_ISL_68191   ····················  ····················  ·····················  ····················  ····················  ····················
U   1232 hCoV-19/Ireland/MH-NVRL-20G47381/2020|EPI_ISL_6819   ····················  ····················  ·····················  ····················  ····················  ····················
U   1233 hCoV-19/Ireland/MH-NVRL-20G47917/2020|EPI_ISL_6819   ····················  ····················  ·····················  ····················  ····················  ····················
U   1234 hCoV-19/Ireland/KE-NVRL-20G47387/2020|EPI_ISL_6819   ····················  ····················  ·····················  ····················  ····················  ····················
U   1235 hCoV-19/Ireland/MH-NVRL-20G49748/2020|EPI_ISL_6819   ····················  ····················  ·····················  ····················  ····················  ····················
U   1236 hCoV-19/Ireland/D-NVRL-20G44042/2020|EPI_ISL_68191   ····················  ····················  ·····················  ····················  ····················  ····················
U   1237 hCoV-19/Ireland/LS-NVRL-20G44005/2020|EPI_ISL_6819   ····················  ····················  ·····················  ····················  ····················  ····················
U   1238 hCoV-19/Ireland/CO-NVRL-20G44155/2020|EPI_ISL_6819   ····················  ·A··················  ·····················  ····················  ····················  ····················
U   1239 hCoV-19/Ireland/D-NVRL-20G41451/2020|EPI_ISL_68192   ····················  ····················  ·····················  ····················  ····················  ····················
U   1240 hCoV-19/Ireland/CO-NVRL-20G44154/2020|EPI_ISL_6819   ····················  ····················  ·····················  ····················  ····················  ····················
U   1241 hCoV-19/Ireland/WH-NVRL-20G49956/2020|EPI_ISL_6819   ····················  ····················  ·····················  ····················  ····················  ····················
U   1242 hCoV-19/Ireland/D-NVRL-20G43510/2020|EPI_ISL_68192   ····················  ····················  ·····················  ····················  ····················  ····················
U   1243 hCoV-19/Ireland/MH-NVRL-20G49952/2020|EPI_ISL_6819   ····················  ····················  ·····················  ····················  ····················  ····················
U   1244 hCoV-19/Ireland/WH-NVRL-20G49966/2020|EPI_ISL_6819   ····················  ····················  ·····················  ····················  ····················  ····················
U   1245 hCoV-19/Ireland/KK-NVRL-20G43857/2020|EPI_ISL_6819   ····················  ····················  ·····················  ····················  ····················  ····················
U   1246 hCoV-19/USA/PA-MGEL-00001/2020|EPI_ISL_681929|2020   ····················  ····················  ·····················  ····················  ····················  ····················
U   1247 hCoV-19/USA/PA-MGEL-00442/2020|EPI_ISL_681930|2020   ····················  ····················  ·····················  ····················  ····················  ····················
U   1248 hCoV-19/USA/PA-MGEL-00443/2020|EPI_ISL_681931|2020   ····················  ····················  ·····················  ····················  ····················  ····················
U   1249 hCoV-19/USA/PA-MGEL-00444/2020|EPI_ISL_681932|2020   ····················  ····················  ·····················  ····················  ····················  ····················
U   1250 hCoV-19/USA/PA-MGEL-00445/2020|EPI_ISL_681933|2020   ····················  ····················  ·····················  ····················  ····················  ····················
U   1251 hCoV-19/USA/PA-MGEL-00446/2020|EPI_ISL_681934|2020   ····················  ····················  ·····················  ····················  ····················  ····················
U   1252 hCoV-19/USA/PA-MGEL-00447/2020|EPI_ISL_681935|2020   ····················  ····················  ·····················  ····················  ····················  ····················
U   1253 hCoV-19/USA/PA-MGEL-00708/2020|EPI_ISL_681936|2020   ····················  ·A··················  ·····················  ····················  ····················  ····················
U   1254 hCoV-19/USA/PA-MGEL-00674/2020|EPI_ISL_681937|2020   ····················  ····················  ·····················  ····················  ····················  ····················
U   1255 hCoV-19/USA/PA-MGEL-00210/2020|EPI_ISL_681939|2020   ····················  ····················  ·····················  ····················  ····················  ····················
U   1256 hCoV-19/USA/PA-MGEL-00002/2020|EPI_ISL_681940|2020   ····················  ····················  ·····················  ····················  ····················  ····················
U   1257 hCoV-19/USA/PA-MGEL-00003/2020|EPI_ISL_681941|2020   ····················  ····················  ·····················  ····················  ····················  ····················
U   1258 hCoV-19/USA/PA-MGEL-00004/2020|EPI_ISL_681942|2020   ····················  ····················  ·····················  ····················  ····················  ····················
U   1259 hCoV-19/USA/PA-MGEL-00005/2020|EPI_ISL_681943|2020   ····················  ····················  ·····················  ····················  ····················  ····················
U   1260 hCoV-19/USA/PA-MGEL-00006/2020|EPI_ISL_681944|2020   ····················  ····················  ·····················  ····················  ····················  ····················
U   1261 hCoV-19/USA/PA-MGEL-00007/2020|EPI_ISL_681945|2020   ····················  ····················  ·····················  ····················  ····················  ····················
U   1262 hCoV-19/USA/PA-MGEL-00019/2020|EPI_ISL_681946|2020   ····················  ····················  ·····················  ····················  ····················  ····················
U   1263 hCoV-19/USA/PA-MGEL-00156/2020|EPI_ISL_681947|2020   ····················  ····················  ·····················  ····················  ····················  ····················
U   1264 hCoV-19/USA/PA-MGEL-00158/2020|EPI_ISL_681948|2020   ····················  ····················  ·····················  ····················  ····················  ····················
U   1265 hCoV-19/USA/PA-MGEL-00159/2020|EPI_ISL_681949|2020   ····················  ····················  ·····················  ····················  ····················  ····················
U   1266 hCoV-19/USA/PA-MGEL-00160/2020|EPI_ISL_681950|2020   ····················  ····················  ·····················  ····················  ····················  ····················
U   1267 hCoV-19/USA/PA-MGEL-00162/2020|EPI_ISL_681951|2020   ····················  ····················  ·····················  ····················  ····················  ····················
U   1268 hCoV-19/USA/PA-MGEL-00163/2020|EPI_ISL_681952|2020   ····················  ····················  ·····················  ····················  ····················  ····················
U   1269 hCoV-19/USA/PA-MGEL-00165/2020|EPI_ISL_681953|2020   ····················  ····················  ·····················  ····················  ····················  ····················
U   1270 hCoV-19/USA/PA-MGEL-00166/2020|EPI_ISL_681954|2020   ····················  ····················  ·····················  ····················  ····················  ····················
U   1271 hCoV-19/USA/PA-MGEL-00257/2020|EPI_ISL_681990|2020   ····················  ····················  ·····················  ····················  ····················  ····················
U   1272 hCoV-19/USA/PA-MGEL-00260/2020|EPI_ISL_681991|2020   ····················  ····················  ·····················  ····················  ····················  ····················
U   1273 hCoV-19/USA/PA-MGEL-00262/2020|EPI_ISL_681992|2020   ····················  ····················  ·····················  ····················  ····················  ····················
U   1274 hCoV-19/USA/PA-MGEL-00263/2020|EPI_ISL_681993|2020   ····················  ····················  ·····················  ····················  ····················  ····················
U   1275 hCoV-19/USA/PA-MGEL-00264/2020|EPI_ISL_681994|2020   ····················  ····················  ·····················  ····················  ····················  ····················
U   1276 hCoV-19/USA/PA-MGEL-00272/2020|EPI_ISL_681995|2020   ····················  ····················  ·····················  ····················  ····················  ····················
U   1277 hCoV-19/USA/PA-MGEL-00273/2020|EPI_ISL_681996|2020   ····················  ····················  ·····················  ····················  ····················  ····················
U   1278 hCoV-19/USA/PA-MGEL-00274/2020|EPI_ISL_681997|2020   ····················  ····················  ·····················  ····················  ····················  ····················
U   1279 hCoV-19/USA/PA-MGEL-00276/2020|EPI_ISL_681998|2020   ····················  ····················  ·····················  ····················  ····················  ····················
U   1280 hCoV-19/USA/PA-MGEL-00324/2020|EPI_ISL_681999|2020   ····················  ····················  ·····················  ····················  ····················  ····················
U   1281 hCoV-19/England/CAMC-B9A45F/2020|EPI_ISL_673400|20   ····················  ····················  ·····················  ····················  ····················  ····················
U   1282 hCoV-19/England/CAMC-B9A325/2020|EPI_ISL_673402|20   ····················  ····················  ·····················  ····················  ····················  ····················
U   1283 hCoV-19/England/CAMC-B9A10D/2020|EPI_ISL_673404|20   ····················  ····················  ·····················  ····················  ····················  ····················
U   1284 hCoV-19/England/CAMC-B99B00/2020|EPI_ISL_673405|20   ····················  ····················  ·····················  ····················  ····················  ····················
U   1285 hCoV-19/England/CAMC-B9ADEB/2020|EPI_ISL_673408|20   ····················  ····················  ·····················  ····················  ····················  ····················
U   1286 hCoV-19/England/CAMC-B9AF9A/2020|EPI_ISL_673409|20   ····················  ····················  ·····················  ····················  ······T·············  ····················
U   1287 hCoV-19/England/CAMC-B9AA6C/2020|EPI_ISL_673410|20   ····················  ····················  ·····················  ····················  ····················  ····················
U   1288 hCoV-19/England/CAMC-B994F6/2020|EPI_ISL_673411|20   ····················  ····················  ·····················  ····················  ····················  ····················
U   1289 hCoV-19/England/CAMC-B99414/2020|EPI_ISL_673412|20   ····················  ····················  ·····················  ····················  ····················  ····················
U   1290 hCoV-19/England/CAMC-B991E0/2020|EPI_ISL_673413|20   ····················  ····················  ·····················  ····················  ····················  ····················
U   1291 hCoV-19/England/CAMC-B99265/2020|EPI_ISL_673415|20   ····················  ····················  ·····················  ····················  ····················  ····················
U   1292 hCoV-19/England/CAMC-B99687/2020|EPI_ISL_673416|20   ····················  ····················  ·····················  ····················  ····················  ····················
U   1293 hCoV-19/England/CAMC-B99E16/2020|EPI_ISL_673417|20   ····················  ····················  ·····················  ····················  ····················  ····················
U   1294 hCoV-19/England/CAMC-B9AA20/2020|EPI_ISL_673420|20   ····················  ····················  ·····················  ····················  ····················  ····················
U   1295 hCoV-19/England/CAMC-B9C4E4/2020|EPI_ISL_673421|20   ····················  ····················  ·····················  ····················  ····················  ····················
U   1296 hCoV-19/England/CAMC-B9A431/2020|EPI_ISL_673436|20   ····················  ····················  ·····················  ····················  ····················  ····················
U   1297 hCoV-19/England/CAMC-B9A2CE/2020|EPI_ISL_673437|20   ····················  ····················  ·····················  ····················  ····················  ····················
U   1298 hCoV-19/England/CAMC-B995D5/2020|EPI_ISL_673438|20   ····················  ····················  ·····················  ····················  ····················  ····················
U   1299 hCoV-19/England/CAMC-B9907A/2020|EPI_ISL_673439|20   ····················  ····················  ·····················  ····················  ····················  ····················
U   1300 hCoV-19/England/CAMC-B99256/2020|EPI_ISL_673445|20   ····················  ····················  ·····················  ····················  ····················  ····················
U   1301 hCoV-19/England/CAMC-B9C5C3/2020|EPI_ISL_673446|20   ····················  ····················  ·····················  ····················  ······T·············  ····················
```

```
U  1302 hCoV-19/England/CAMC-B9ADFA/2020|EPI_ISL_673448|20  ···················· ·A·················· ···················· ···················· ···················· ····················
U  1303 hCoV-19/England/CAMC-B9A04C/2020|EPI_ISL_673449|20  ···················· ···················· ···················· ···················· ···················· ····················
U  1304 hCoV-19/England/CAMC-B99784/2020|EPI_ISL_673450|20  ···················· ···················· ···················· ···················· ···················· ····················
U  1305 hCoV-19/England/CAMC-B993AE/2020|EPI_ISL_673451|20  ···················· ···················· ···················· ···················· ···················· ····················
U  1306 hCoV-19/England/CAMC-B99335/2020|EPI_ISL_673453|20  ···················· ···················· ···················· ···················· ···················· ····················
U  1307 hCoV-19/England/CAMC-B98B3E/2020|EPI_ISL_673454|20  ···················· ···················· ···················· ···················· ···················· ····················
U  1308 hCoV-19/England/CAMC-B99D82/2020|EPI_ISL_673455|20  ···················· ···················· ···················· ···················· ···················· ····················
U  1309 hCoV-19/England/CAMC-B99089/2020|EPI_ISL_673456|20  ········T··········· ···················· ···················· ···················· ···················· ····················
U  1310 hCoV-19/England/CAMC-B99C49/2020|EPI_ISL_673457|20  ···················· ···················· ···················· ···················· ···················· ····················
U  1311 hCoV-19/England/CAMC-B9AA02/2020|EPI_ISL_673458|20  ···················· ···················· ···················· ···················· ···················· ····················
U  1312 hCoV-19/England/CAMC-B99441/2020|EPI_ISL_673459|20  ···················· ···················· ···················· ···················· ···················· ····················
U  1313 hCoV-19/England/CAMC-B9A0A6/2020|EPI_ISL_673460|20  ···················· ···················· ···················· ···················· ···················· ····················
U  1314 hCoV-19/England/CAMC-B99BA6/2020|EPI_ISL_673461|20  ···················· ···················· ···················· ···················· ···················· ····················
U  1315 hCoV-19/England/CAMC-B9A3BC/2020|EPI_ISL_673462|20  ···················· ···················· ···················· ···················· ···················· ····················
U  1316 hCoV-19/England/CAMC-B98F41/2020|EPI_ISL_673463|20  ···················· ···················· ···················· ···················· ······T············· ····················
U  1317 hCoV-19/England/CAMC-B99AC7/2020|EPI_ISL_673464|20  ···················· ···················· ···················· ···················· ···················· ····················
U  1318 hCoV-19/England/CAMC-B9A12B/2020|EPI_ISL_673465|20  ···················· ···················· ···················· ···················· ···················· ····················
U  1319 hCoV-19/England/CAMC-B997FD/2020|EPI_ISL_673467|20  ···················· ···················· ···················· ···················· ···················· ····················
U  1320 hCoV-19/England/CAMC-B99247/2020|EPI_ISL_673468|20  ···················· ···················· ···················· ···················· ···················· ····················
U  1321 hCoV-19/England/CAMC-B992ED/2020|EPI_ISL_673469|20  ···················· ···················· ···················· ···················· ···················· ····················
U  1322 hCoV-19/England/CAMC-B9A185/2020|EPI_ISL_673470|20  ···················· ···················· ···················· ···················· ···················· ····················
U  1323 hCoV-19/England/CAMC-B99CFE/2020|EPI_ISL_673471|20  ···················· ···················· ···················· ···················· ···················· ····················
U  1324 hCoV-19/England/CAMC-B99292/2020|EPI_ISL_673473|20  ···················· ···················· ···················· ···················· ···················· ····················
U  1325 hCoV-19/England/CAMC-B99E43/2020|EPI_ISL_673474|20  ···················· ···················· ···················· ···················· ···················· ····················
U  1326 hCoV-19/England/CAMC-B9C578/2020|EPI_ISL_673476|20  ···················· ···················· ···················· ···················· ···················· ····················
U  1327 hCoV-19/England/CAMC-B98B4D/2020|EPI_ISL_673478|20  ···················· ···················· ···················· ···················· ···················· ····················
U  1328 hCoV-19/England/CAMC-B99317/2020|EPI_ISL_673479|20  ···················· ···················· ···················· ···················· ···················· ····················
U  1329 hCoV-19/England/CAMC-B9C60C/2020|EPI_ISL_673480|20  ···················· ···················· ···················· ···················· ···················· ····················
U  1330 hCoV-19/England/CAMC-B99D91/2020|EPI_ISL_673482|20  ···················· ···················· ···················· ···················· ···················· ····················
U  1331 hCoV-19/England/CAMC-B9A52F/2020|EPI_ISL_673483|20  ···················· ···················· ···················· ···················· ···················· ····················
U  1332 hCoV-19/England/CAMC-B99405/2020|EPI_ISL_673485|20  ···················· ···················· ···················· ···················· ···················· ····················
U  1333 hCoV-19/England/CAMC-B9A97E/2020|EPI_ISL_673486|20  ···················· ···················· ···················· ···················· ···················· ····················
U  1334 hCoV-19/England/CAMC-B9A4E6/2020|EPI_ISL_673487|20  ···················· ···················· ···················· ···················· ···················· ····················
U  1335 hCoV-19/England/CAMC-B9A02E/2020|EPI_ISL_673488|20  ···················· ···················· ···················· ···················· ···················· ····················
U  1336 hCoV-19/England/CAMC-B99669/2020|EPI_ISL_673489|20  ···················· ···················· ···················· ···················· ···················· ····················
U  1337 hCoV-19/England/CAMC-B9C51E/2020|EPI_ISL_673491|20  ···················· ·A·················· ···················· ···················· ···················· ···A················
U  1338 hCoV-19/England/CAMC-B992DE/2020|EPI_ISL_673493|20  ···················· ···················· ···················· ···················· ···················· ····················
U  1339 hCoV-19/England/CAMC-B99AF4/2020|EPI_ISL_673494|20  ···················· ···················· ···················· ···················· ···················· ····················
U  1340 hCoV-19/England/CAMC-B99C85/2020|EPI_ISL_673495|20  ···················· ···················· ···················· ···················· ···················· ····················
U  1341 hCoV-19/England/CAMC-B9A05B/2020|EPI_ISL_673496|20  ···················· ···················· ···················· ···················· ···················· ····················
U  1342 hCoV-19/England/CAMC-B997A2/2020|EPI_ISL_673497|20  ···················· ···················· ···················· ···················· ···················· ····················
U  1343 hCoV-19/England/CAMC-B9AC39/2020|EPI_ISL_673498|20  ···················· ···················· ···················· ···················· ···················· ····················
U  1344 hCoV-19/England/CAMC-B9A4B9/2020|EPI_ISL_673499|20  ···················· ···················· ···················· ···················· ···················· ····················
U  1345 hCoV-19/England/MILK-B8EA7A/2020|EPI_ISL_674700|20  ···················· ···················· ···················· ···················· ···················· ····················
U  1346 hCoV-19/England/MILK-B8EAD4/2020|EPI_ISL_674701|20  ···················· ···················· ···················· ···················· ······T············· ····················
U  1347 hCoV-19/England/MILK-B8F88E/2020|EPI_ISL_674702|20  ···················· ···················· ···················· ···················· ···················· ····················
U  1348 hCoV-19/England/MILK-B8F50F/2020|EPI_ISL_674704|20  ···················· ···················· ···················· ···················· ···················· ····················
U  1349 hCoV-19/England/MILK-B8F8BB/2020|EPI_ISL_674705|20  ···················· ···················· ···················· ···················· ···················· ····················
U  1350 hCoV-19/England/MILK-B8F47B/2020|EPI_ISL_674706|20  ···················· ···················· ···················· ···················· ···················· ····················
U  1351 hCoV-19/England/QEUH-B7CECE/2020|EPI_ISL_674707|20  ···················· ···················· ···················· ···················· ···················· ····················
U  1352 hCoV-19/England/MILK-B8FA00/2020|EPI_ISL_674708|20  ···················· ···················· ···················· ···················· ···················· ····················
U  1353 hCoV-19/England/MILK-B8ECB0/2020|EPI_ISL_674709|20  ···················· ···················· ···················· ···················· ···················· ····················
U  1354 hCoV-19/England/MILK-B8F6FD/2020|EPI_ISL_674710|20  ···················· ···················· ···················· ···················· ···················· ····················
U  1355 hCoV-19/England/MILK-B8FA5B/2020|EPI_ISL_674712|20  ···················· ···················· ···················· ···················· ···················· ····················
U  1356 hCoV-19/England/QEUH-B7CF9E/2020|EPI_ISL_674714|20  ···················· ···················· ···················· ···················· ···················· ····················
U  1357 hCoV-19/England/QEUH-B7D6E3/2020|EPI_ISL_674716|20  ···················· ···················· ···················· ···················· ···················· ····················
U  1358 hCoV-19/England/QEUH-B7CE46/2020|EPI_ISL_674717|20  ···················· ···················· ···················· ···················· ···················· ····················
U  1359 hCoV-19/England/MILK-B8E8E9/2020|EPI_ISL_674719|20  ···················· ···················· ···················· ···················· ···················· ····················
U  1360 hCoV-19/England/MILK-B8EB3B/2020|EPI_ISL_674720|20  ···················· ···················· ···················· ···················· ···················· ····················
U  1361 hCoV-19/England/MILK-B8DDEB/2020|EPI_ISL_674721|20  ···················· ···················· ···················· ···················· ···················· ····················
U  1362 hCoV-19/England/QEUH-B7F58A/2020|EPI_ISL_674723|20  ···················· ···················· ···················· ···················· ···················· ····················
U  1363 hCoV-19/England/QEUH-B7F48D/2020|EPI_ISL_674724|20  ···················· ···················· ···················· ···················· ···················· ····················
U  1364 hCoV-19/England/QEUH-B7F4AB/2020|EPI_ISL_674725|20  ···················· ···················· ···················· ···················· ···················· ····················
U  1365 hCoV-19/England/MILK-B8DEAC/2020|EPI_ISL_674728|20  ···················· ···················· ···················· ···················· ···················· ····················
U  1366 hCoV-19/England/MILK-B8E096/2020|EPI_ISL_674729|20  ···················· ·A·················· ···················· ···················· ···················· ···A················
U  1367 hCoV-19/England/MILK-B8EBE0/2020|EPI_ISL_674730|20  ···················· ···················· ···················· ···················· ···················· ····················
U  1368 hCoV-19/England/MILK-B8DE7F/2020|EPI_ISL_674731|20  ···················· ···················· ···················· ···················· ···················· ····················
U  1369 hCoV-19/England/MILK-B8F903/2020|EPI_ISL_674732|20  ···················· ···················· ···················· ···················· ···················· ····················
U  1370 hCoV-19/England/MILK-B8E7EC/2020|EPI_ISL_674733|20  ···················· ···················· ···················· ···················· ···················· ····················
U  1371 hCoV-19/England/MILK-B8DD72/2020|EPI_ISL_674734|20  ···················· ···················· ···················· ···················· ···················· ····················
U  1372 hCoV-19/England/MILK-B8DF6D/2020|EPI_ISL_674735|20  ···················· ···················· ···················· ···················· ······T············· ····················
U  1373 hCoV-19/England/QEUH-B8097B/2020|EPI_ISL_674736|20  ···················· ···················· ···················· ···················· ···················· ····················
U  1374 hCoV-19/England/MILK-B8E8F8/2020|EPI_ISL_674737|20  ···················· ···················· ···················· ···················· ···················· ····················
U  1375 hCoV-19/England/MILK-B8EB77/2020|EPI_ISL_674738|20  ···················· ···················· ···················· ···················· ···················· ····················
U  1376 hCoV-19/England/MILK-B8DDFA/2020|EPI_ISL_674739|20  ···················· ···················· ···················· ···················· ···················· ····················
U  1377 hCoV-19/England/MILK-B8F499/2020|EPI_ISL_674742|20  ···················· ···················· ···················· ···················· ···················· ····················
U  1378 hCoV-19/England/QEUH-B7CE64/2020|EPI_ISL_674743|20  ···················· ···················· ···················· ···················· ···················· ····················
U  1379 hCoV-19/England/QEUH-B808C9/2020|EPI_ISL_674744|20  ···················· ···················· ···················· ···················· ···················· ····················
U  1380 hCoV-19/England/QEUH-B808D8/2020|EPI_ISL_674745|20  ···················· ···················· ···················· ···················· ···················· ····················
U  1381 hCoV-19/England/QEUH-B7D65C/2020|EPI_ISL_674746|20  ···················· ···················· ···················· ···················· ···················· ····················
```

| | | | | | | | | |
|---|---|---|---|---|---|---|---|---|
| U | 1382 | hCoV-19/England/QEUH-B7F8BE/2020\|EPI_ISL_674747\|20 | ···················· | ···················· | ····················· | ···················· | ···················· | ···················· |
| U | 1383 | hCoV-19/England/MILK-B8F772/2020\|EPI_ISL_674748\|20 | ···················· | ···················· | ····················· | ···················· | ···················· | ···················· |
| U | 1384 | hCoV-19/England/QEUH-B7F809/2020\|EPI_ISL_674749\|20 | ···················· | ···················· | ····················· | ···················· | ···················· | ···················· |
| U | 1385 | hCoV-19/England/MILK-B8EB4A/2020\|EPI_ISL_674750\|20 | ···················· | ···················· | ····················· | ···················· | ···················· | ···················· |
| U | 1386 | hCoV-19/England/QEUH-B7F5A8/2020\|EPI_ISL_674751\|20 | ···················· | ···················· | ····················· | ···················· | ···················· | ···················· |
| U | 1387 | hCoV-19/England/MILK-B8E0C3/2020\|EPI_ISL_674764\|20 | ···················· | ···················· | ····················· | ···················· | ···················· | ···················· |
| U | 1388 | hCoV-19/England/MILK-B8E44F/2020\|EPI_ISL_674765\|20 | ···················· | ···················· | ····················· | ···················· | ···················· | ···················· |
| U | 1389 | hCoV-19/England/MILK-B8EA2F/2020\|EPI_ISL_674766\|20 | ···················· | ···················· | ····················· | ···················· | ···················· | ···················· |
| U | 1390 | hCoV-19/England/MILK-B8F833/2020\|EPI_ISL_674767\|20 | ···················· | ···················· | ····················· | ···················· | ···················· | ···················· |
| U | 1391 | hCoV-19/England/MILK-B8E9C8/2020\|EPI_ISL_674768\|20 | ···················· | ···················· | ····················· | ···················· | ···················· | ···················· |
| U | 1392 | hCoV-19/England/MILK-B8F860/2020\|EPI_ISL_674769\|20 | ···················· | ···················· | ····················· | ···················· | ···················· | ···················· |
| U | 1393 | hCoV-19/England/MILK-B8EC65/2020\|EPI_ISL_674777\|20 | ···················· | ···················· | ····················· | ···················· | ···················· | ···················· |
| U | 1394 | hCoV-19/England/MILK-B8ECFC/2020\|EPI_ISL_674778\|20 | ···················· | ···················· | ····················· | ···················· | ······T············· | ···················· |
| U | 1395 | hCoV-19/England/QEUH-B7F757/2020\|EPI_ISL_674779\|20 | ···················· | ···················· | ····················· | ···················· | ···················· | ···················· |
| U | 1396 | hCoV-19/England/QEUH-B7F4BA/2020\|EPI_ISL_674781\|20 | ···················· | ·A·················· | ····················· | ···················· | ···················· | ···················· |
| U | 1397 | hCoV-19/England/QEUH-B7D610/2020\|EPI_ISL_674783\|20 | ···················· | ···················· | ····················· | ···················· | ···················· | ···················· |
| U | 1398 | hCoV-19/England/MILK-B8F930/2020\|EPI_ISL_674784\|20 | ···················· | ···················· | ····················· | ···················· | ···················· | ···················· |
| U | 1399 | hCoV-19/England/MILK-B8F7CD/2020\|EPI_ISL_674785\|20 | ···················· | ···················· | ····················· | ···················· | ···················· | ···················· |
| U | 1400 | hCoV-19/England/QEUH-B7CE55/2020\|EPI_ISL_674786\|20 | ···················· | ···················· | ····················· | ···················· | ···················· | ···················· |
| U | 1401 | hCoV-19/England/MILK-B8EBA4/2020\|EPI_ISL_674787\|20 | ···················· | ···················· | ····················· | ···················· | ···················· | ···················· |
| U | 1402 | hCoV-19/England/MILK-B8E46D/2020\|EPI_ISL_674788\|20 | ···················· | ···················· | ····················· | ···················· | ···················· | ···················· |
| U | 1403 | hCoV-19/England/MILK-B8DBF0/2020\|EPI_ISL_674789\|20 | ···················· | ···················· | ····················· | ···················· | ···················· | ···················· |
| U | 1404 | hCoV-19/England/MILK-B8E4C7/2020\|EPI_ISL_674790\|20 | ···················· | ···················· | ····················· | ···················· | ···················· | ···················· |
| U | 1405 | hCoV-19/England/QEUH-B7F5C6/2020\|EPI_ISL_674791\|20 | ···················· | ···················· | ····················· | ···················· | ···················· | ···················· |
| U | 1406 | hCoV-19/England/QEUH-B8098A/2020\|EPI_ISL_674792\|20 | ···················· | ···················· | ····················· | ···················· | ···················· | ···················· |
| U | 1407 | hCoV-19/England/MILK-B8EB59/2020\|EPI_ISL_674793\|20 | ···················· | ···················· | ····················· | ···················· | ···················· | ···················· |
| U | 1408 | hCoV-19/England/QEUH-B7D6C5/2020\|EPI_ISL_674794\|20 | ···················· | ···················· | ····················· | ···················· | ···················· | ···················· |
| U | 1409 | hCoV-19/England/MILK-B8F5A5/2020\|EPI_ISL_674796\|20 | ···················· | ···················· | ····················· | ···················· | ···················· | ···················· |
| U | 1410 | hCoV-19/England/MILK-B8E98C/2020\|EPI_ISL_674797\|20 | ···················· | ···················· | ····················· | ···················· | ···················· | ···················· |
| U | 1411 | hCoV-19/England/MILK-B8DD63/2020\|EPI_ISL_674798\|20 | ···················· | ···················· | ····················· | ···················· | ···················· | ···················· |
| U | 1412 | hCoV-19/England/MILK-B8E39D/2020\|EPI_ISL_674799\|20 | ···················· | ···················· | ····················· | ···················· | ···················· | ···················· |
| U | 1413 | hCoV-19/USA/NY-Wadsworth-192868-01/2020\|EPI_ISL_67 | ···················· | ··············C····· | ····················· | ···················· | ···················· | ···················· |
| U | 1414 | hCoV-19/USA/NY-Wadsworth-192870-01/2020\|EPI_ISL_67 | ···················· | ···················· | ····················· | ···················· | ···················· | ···················· |
| U | 1415 | hCoV-19/USA/NY-Wadsworth-192871-01/2020\|EPI_ISL_67 | ···················· | ···················· | ····················· | ···················· | ···················· | ···················· |
| U | 1416 | hCoV-19/USA/NY-Wadsworth-192872-01/2020\|EPI_ISL_67 | ···················· | ···················· | ····················· | ···················· | ···················· | ···················· |
| U | 1417 | hCoV-19/USA/NY-Wadsworth-192873-01/2020\|EPI_ISL_67 | ···················· | ···················· | ····················· | ···················· | ···················· | ···················· |
| U | 1418 | hCoV-19/USA/NY-Wadsworth-192874-01/2020\|EPI_ISL_67 | ···················· | ··············C····· | ····················· | ···················· | ···················· | ···················· |
| U | 1419 | hCoV-19/USA/NY-Wadsworth-192875-01/2020\|EPI_ISL_67 | ···················· | ···················· | ····················· | ···················· | ···················· | ···················· |
| U | 1420 | hCoV-19/USA/NY-Wadsworth-192876-01/2020\|EPI_ISL_67 | ···················· | ··············C····· | ····················· | ···················· | ···················· | ···················· |
| U | 1421 | hCoV-19/USA/NY-Wadsworth-192877-01/2020\|EPI_ISL_67 | ···················· | ··············C····· | ····················· | ···················· | ···················· | ···················· |
| U | 1422 | hCoV-19/USA/NY-Wadsworth-192879-01/2020\|EPI_ISL_67 | ···················· | ··············C····· | ····················· | ···················· | ···················· | ···················· |
| U | 1423 | hCoV-19/USA/NY-Wadsworth-192889-01/2020\|EPI_ISL_67 | ···················· | ··············C····· | ····················· | ···················· | ···················· | ···················· |
| U | 1424 | hCoV-19/USA/NY-Wadsworth-192890-01/2020\|EPI_ISL_67 | ···················· | ··············C····· | ····················· | ···················· | ···················· | ···················· |
| U | 1425 | hCoV-19/Russia/SAR-RARI-2666/2020\|EPI_ISL_678280\|2 | ···················· | ···················· | ····················· | ···················· | ···················· | ···················· |
| U | 1426 | hCoV-19/Russia/SAR-RARI-3080/2020\|EPI_ISL_678281\|2 | ···················· | ···················· | ····················· | ···················· | ···················· | ···················· |
| U | 1427 | hCoV-19/Russia/SAR-RARI-3656/2020\|EPI_ISL_678282\|2 | ···················· | ···················· | ····················· | ···················· | ···················· | ···················· |
| U | 1428 | hCoV-19/Russia/SAR-RARI-3573/2020\|EPI_ISL_678283\|2 | ···················· | ···················· | ····················· | ···················· | ···················· | ···················· |
| U | 1429 | hCoV-19/Russia/SAR-RARI-2272/2020\|EPI_ISL_678284\|2 | ···················· | ···················· | ····················· | ···················· | ···················· | ···················· |
| U | 1430 | hCoV-19/Russia/SAR-RARI-3083/2020\|EPI_ISL_678285\|2 | ···················· | ···················· | ····················· | ···················· | ···················· | ···················· |
| U | 1431 | hCoV-19/Russia/SAR-RARI-9895/2020\|EPI_ISL_678286\|2 | ···················· | ···················· | ····················· | ···················· | ···················· | ···················· |
| U | 1432 | hCoV-19/Russia/MOS-RARI-940/2020\|EPI_ISL_678287\|20 | ···················· | ···················· | ····················· | ···················· | ···················· | ···················· |
| U | 1433 | hCoV-19/Russia/PEN-RARI-953/2020\|EPI_ISL_678288\|20 | ···················· | ···················· | ····················· | ···················· | ···················· | ···················· |
| U | 1434 | hCoV-19/Hungary/US-60441w-2/2020\|EPI_ISL_678290\|20 | ···················· | ···················· | ····················· | ···················· | ···················· | ···················· |
| U | 1435 | hCoV-19/Hungary/US-60558w/2020\|EPI_ISL_678291\|2020 | ···················· | ···················· | ····················· | ···················· | ···················· | ···················· |
| U | 1436 | hCoV-19/Australia/NSW2028/2020\|EPI_ISL_678293\|2020 | ···················· | ···················· | ····················· | ···················· | ···················· | ···················· |
| U | 1437 | hCoV-19/Australia/NSW2099/2020\|EPI_ISL_678297\|2020 | ···················· | ···················· | ····················· | ···················· | ···················· | ···················· |
| U | 1438 | hCoV-19/Australia/NSW2110/2020\|EPI_ISL_678299\|2020 | ···················· | ···················· | ····················· | ···················· | ···················· | ···················· |
| U | 1439 | hCoV-19/Australia/NSW2589/2020\|EPI_ISL_678318\|2020 | ···················· | ···················· | ····················· | ···················· | ···················· | ···················· |
| U | 1440 | hCoV-19/Australia/NSW2603/2020\|EPI_ISL_678320\|2020 | ···················· | ···················· | ····················· | ···················· | ···················· | ···················· |
| U | 1441 | hCoV-19/Australia/NSW2609/2020\|EPI_ISL_678321\|2020 | ···················· | ···················· | ····················· | ···················· | ···················· | ···················· |
| U | 1442 | hCoV-19/Australia/NSW2624/2020\|EPI_ISL_678322\|2020 | ···················· | ···················· | ····················· | ···················· | ···················· | ···················· |
| U | 1443 | hCoV-19/Australia/NSW2626/2020\|EPI_ISL_678323\|2020 | ···················· | ···················· | ····················· | ···················· | ···················· | ···················· |
| U | 1444 | hCoV-19/Australia/NSW2631/2020\|EPI_ISL_678325\|2020 | ···················· | ···················· | ····················· | ···················· | ···················· | ···················· |
| U | 1445 | hCoV-19/Australia/NSW2632/2020\|EPI_ISL_678326\|2020 | ···················· | ···················· | ····················· | ···················· | ···················· | ···················· |
| U | 1446 | hCoV-19/Australia/NSW2633/2020\|EPI_ISL_678327\|2020 | ···················· | ···················· | ····················· | ···················· | ···················· | ···················· |
| U | 1447 | hCoV-19/Australia/NSW2634/2020\|EPI_ISL_678328\|2020 | ···················· | ···················· | ····················· | ···················· | ···················· | ···················· |
| U | 1448 | hCoV-19/Australia/NSW2646/2020\|EPI_ISL_678330\|2020 | ···················· | ···················· | ····················· | ···················· | ···················· | ···················· |
| U | 1449 | hCoV-19/Australia/NSW2648/2020\|EPI_ISL_678331\|2020 | ···················· | ·A·················· | ····················· | ···················· | ···················· | ···A················ |
| U | 1450 | hCoV-19/Australia/NSW2653/2020\|EPI_ISL_678332\|2020 | ···················· | ···················· | ····················· | ···················· | ···················· | ···················· |
| U | 1451 | hCoV-19/Australia/NSW2656/2020\|EPI_ISL_678333\|2020 | ···················· | ···················· | ····················· | ···················· | ···················· | ···················· |
| U | 1452 | hCoV-19/Australia/NSW2692/2020\|EPI_ISL_678336\|2020 | ···················· | ···················· | ····················· | ···················· | ···················· | ···················· |
| U | 1453 | hCoV-19/Australia/NSW2696/2020\|EPI_ISL_678337\|2020 | ···················· | ···················· | ····················· | ···················· | ···················· | ···················· |
| U | 1454 | hCoV-19/Australia/NSW2796/2020\|EPI_ISL_678348\|2020 | ···················· | ···················· | ····················· | ···················· | ···················· | ···················· |
| U | 1455 | hCoV-19/England/MILK-B39892/2020\|EPI_ISL_679648\|20 | ···················· | ···················· | ····················· | ···················· | ···················· | ···················· |
| U | 1456 | hCoV-19/England/MILK-B398B0/2020\|EPI_ISL_679649\|20 | ···················· | ···················· | ····················· | ···················· | ···················· | ···················· |
| U | 1457 | hCoV-19/England/MILK-B398CF/2020\|EPI_ISL_679650\|20 | ···················· | ···················· | ····················· | ···················· | ···················· | ···················· |
| U | 1458 | hCoV-19/England/MILK-B3999F/2020\|EPI_ISL_679652\|20 | ···················· | ···················· | ····················· | ···················· | ···················· | ···················· |
| U | 1459 | hCoV-19/England/MILK-B399EA/2020\|EPI_ISL_679654\|20 | ···················· | ···················· | ····················· | ···················· | ···················· | ···················· |
| U | 1460 | hCoV-19/England/MILK-B399F9/2020\|EPI_ISL_679655\|20 | ···················· | ···················· | ····················· | ···················· | ···················· | ···················· |
| U | 1461 | hCoV-19/England/MILK-B39A05/2020\|EPI_ISL_679656\|20 | ···················· | ···················· | ····················· | ···················· | ···················· | ···················· |

| | | | | | | | | |
|---|---|---|---|---|---|---|---|---|
| U | 1462 hCoV-19/England/MILK-B39A14/2020\|EPI_ISL_679657\|20 | ···················· | ···················· | ···················· | ···················· | ···················· | ···················· | |
| U | 1463 hCoV-19/England/MILK-B39A23/2020\|EPI_ISL_679658\|20 | ···················· | ···················· | ···················· | ···················· | ···················· | ···················· | |
| U | 1464 hCoV-19/England/MILK-B39A32/2020\|EPI_ISL_679659\|20 | ···················· | ···················· | ···················· | ···················· | ···················· | ···················· | |
| U | 1465 hCoV-19/England/MILK-B39A41/2020\|EPI_ISL_679660\|20 | ···················· | ···················· | ···················· | ···················· | ···················· | ···················· | |
| U | 1466 hCoV-19/England/MILK-B39C2D/2020\|EPI_ISL_679662\|20 | ···················· | ···················· | ···················· | ···················· | ·····T·············· | ···················· | |
| U | 1467 hCoV-19/England/MILK-B39C3C/2020\|EPI_ISL_679663\|20 | ···················· | ···················· | ···················· | ···················· | ···················· | ···················· | |
| U | 1468 hCoV-19/England/MILK-B39C78/2020\|EPI_ISL_679665\|20 | ···················· | ···················· | ···················· | ···················· | ···················· | ···················· | |
| U | 1469 hCoV-19/England/MILK-B39DB1/2020\|EPI_ISL_679666\|20 | ···················· | ···················· | ···················· | ···················· | ···················· | ···················· | |
| U | 1470 hCoV-19/England/MILK-B39DDF/2020\|EPI_ISL_679667\|20 | ···················· | ···················· | ···················· | ···················· | ·····T·············· | ···················· | |
| U | 1471 hCoV-19/England/MILK-B39DEE/2020\|EPI_ISL_679668\|20 | ···················· | ···················· | ···················· | ···················· | ···················· | ···················· | |
| U | 1472 hCoV-19/England/MILK-B39E09/2020\|EPI_ISL_679669\|20 | ···················· | ···················· | ···················· | ···················· | ···················· | ···················· | |
| U | 1473 hCoV-19/England/MILK-B3A0C6/2020\|EPI_ISL_679679\|20 | ···················· | ···················· | ···················· | ···················· | ···················· | ···················· | |
| U | 1474 hCoV-19/Wales/PHWC-48D24D/2020\|EPI_ISL_680594\|2020 | ··················Y· | ···················· | ···················· | ···················· | ···················· | ···················· | 1 |
| U | 1475 hCoV-19/Wales/PHWC-48D25C/2020\|EPI_ISL_680595\|2020 | ···················· | ···················· | ···················· | ···················· | ···················· | ···················· | |
| U | 1476 hCoV-19/Wales/PHWC-48D26B/2020\|EPI_ISL_680596\|2020 | ···················· | ···················· | ···················· | ···················· | ···················· | ···················· | |
| U | 1477 hCoV-19/Wales/PHWC-48D27A/2020\|EPI_ISL_680597\|2020 | ···················· | ···················· | ···················· | ···················· | ···················· | ···················· | |
| U | 1478 hCoV-19/Wales/PHWC-48D289/2020\|EPI_ISL_680598\|2020 | ···················· | ···················· | ···················· | ···················· | ···················· | ···················· | |
| U | 1479 hCoV-19/Wales/PHWC-48D298/2020\|EPI_ISL_680599\|2020 | ···················· | ···················· | ···················· | ···················· | ···················· | ···················· | |
| U | 1480 hCoV-19/Wales/PHWC-48D492/2020\|EPI_ISL_680630\|2020 | ···················· | ···················· | ···················· | ···················· | ···················· | ···················· | |
| U | 1482 hCoV-19/Wales/PHWC-48D4B0/2020\|EPI_ISL_680632\|2020 | ···················· | ···················· | ···················· | ···················· | ···················· | ···················· | |
| U | 1483 hCoV-19/Wales/PHWC-48D4CF/2020\|EPI_ISL_680633\|2020 | ···················· | ···················· | ···················· | ···················· | ···················· | ···················· | |
| U | 1484 hCoV-19/Wales/PHWC-48D4DE/2020\|EPI_ISL_680634\|2020 | ···················· | ···················· | ···················· | ···················· | ···················· | ···················· | |
| U | 1485 hCoV-19/Wales/PHWC-48D4ED/2020\|EPI_ISL_680635\|2020 | ···················· | ···················· | ···················· | ···················· | ···················· | ···················· | |
| U | 1486 hCoV-19/Wales/PHWC-48D4FC/2020\|EPI_ISL_680636\|2020 | ···················· | ···················· | ···················· | ···················· | ···················· | ···················· | |
| U | 1487 hCoV-19/Wales/PHWC-48D508/2020\|EPI_ISL_680637\|2020 | ···················· | ···················· | ···················· | ···················· | ···················· | ···················· | |
| U | 1488 hCoV-19/Wales/PHWC-48D526/2020\|EPI_ISL_680639\|2020 | ···················· | ···················· | ···················· | ···················· | ···················· | ···················· | |
| U | 1489 hCoV-19/Wales/PHWC-48D535/2020\|EPI_ISL_680640\|2020 | ···················· | ···················· | ···················· | ···················· | ···················· | ···················· | |
| U | 1490 hCoV-19/Wales/PHWC-48D544/2020\|EPI_ISL_680641\|2020 | ···················· | ···················· | ···················· | ···················· | ···················· | ···················· | |
| U | 1491 hCoV-19/Wales/PHWC-48D553/2020\|EPI_ISL_680642\|2020 | ···················· | ···················· | ···················· | ···················· | ···················· | ···················· | |
| U | 1492 hCoV-19/Wales/PHWC-48D571/2020\|EPI_ISL_680644\|2020 | ···················· | ···················· | ···················· | ···················· | ···················· | ···················· | |
| U | 1493 hCoV-19/Wales/PHWC-48D580/2020\|EPI_ISL_680645\|2020 | ···················· | ···················· | ···················· | ···················· | ···················· | ···················· | |
| U | 1494 hCoV-19/Wales/PHWC-48D5AE/2020\|EPI_ISL_680647\|2020 | ···················· | ···················· | ···················· | ···················· | ···················· | ···················· | |
| U | 1495 hCoV-19/Wales/PHWC-48D5BD/2020\|EPI_ISL_680648\|2020 | ···················· | ···················· | ···················· | ···················· | ···················· | ···················· | |
| U | 1496 hCoV-19/Wales/PHWC-48D5CC/2020\|EPI_ISL_680649\|2020 | ···················· | ···················· | ···················· | ···················· | ···················· | ···················· | |
| U | 1497 hCoV-19/Wales/PHWC-48D5DB/2020\|EPI_ISL_680650\|2020 | ···················· | ···················· | ···················· | ···················· | ···················· | ···················· | |
| U | 1498 hCoV-19/Wales/PHWC-48D5EA/2020\|EPI_ISL_680651\|2020 | ···················· | ···················· | ···················· | ···················· | ···················· | ···················· | |
| U | 1499 hCoV-19/Wales/PHWC-48D605/2020\|EPI_ISL_680652\|2020 | ···················· | ···················· | ···················· | ···················· | ···················· | ···················· | |
| U | 1500 hCoV-19/Wales/PHWC-48D614/2020\|EPI_ISL_680653\|2020 | ···················· | ···················· | ···················· | ···················· | ···················· | ···················· | |
| U | 1501 hCoV-19/Wales/PHWC-48D623/2020\|EPI_ISL_680654\|2020 | ···················· | ···················· | ···················· | ···················· | ···················· | ···················· | |
| U | 1502 hCoV-19/Wales/PHWC-48D632/2020\|EPI_ISL_680655\|2020 | ···················· | ···················· | ···················· | ···················· | ···················· | ···················· | |
| U | 1503 hCoV-19/Wales/PHWC-48D641/2020\|EPI_ISL_680656\|2020 | ···················· | ···················· | ···················· | ···················· | ···················· | ···················· | |
| U | 1504 hCoV-19/Wales/PHWC-48D650/2020\|EPI_ISL_680657\|2020 | ···················· | ···················· | ···················· | ···················· | ···················· | ···················· | |
| U | 1505 hCoV-19/Wales/PHWC-48D66F/2020\|EPI_ISL_680658\|2020 | ···················· | ···················· | ···················· | ···················· | ···················· | ···················· | |
| U | 1506 hCoV-19/Wales/PHWC-48D67E/2020\|EPI_ISL_680659\|2020 | ···················· | ···················· | ···················· | ···················· | ···················· | ···················· | |
| U | 1507 hCoV-19/USA/PA-MGEL-00204/2020\|EPI_ISL_681960\|2020 | ···················· | ···················· | ···················· | ···················· | ···················· | ···················· | |
| U | 1508 hCoV-19/USA/PA-MGEL-00206/2020\|EPI_ISL_681961\|2020 | ···················· | ···················· | ···················· | ···················· | ···················· | ···················· | |
| U | 1509 hCoV-19/USA/PA-MGEL-00207/2020\|EPI_ISL_681962\|2020 | ···················· | ···················· | ···················· | ···················· | ···················· | ···················· | |
| U | 1510 hCoV-19/USA/PA-MGEL-00208/2020\|EPI_ISL_681963\|2020 | ···················· | ···················· | ···················· | ···················· | ···················· | ···················· | |
| U | 1511 hCoV-19/USA/PA-MGEL-00213/2020\|EPI_ISL_681964\|2020 | ···················· | ···················· | ···················· | ···················· | ···················· | ···················· | |
| U | 1512 hCoV-19/USA/PA-MGEL-00215/2020\|EPI_ISL_681965\|2020 | ···················· | ···················· | ···················· | ···················· | ···················· | ···················· | |
| U | 1513 hCoV-19/USA/PA-MGEL-00216/2020\|EPI_ISL_681966\|2020 | ···················· | ···················· | ···················· | ···················· | ···················· | ···················· | |
| U | 1514 hCoV-19/USA/PA-MGEL-00219/2020\|EPI_ISL_681967\|2020 | ···················· | ···················· | ···················· | ···················· | ···················· | ···················· | |
| U | 1515 hCoV-19/USA/PA-MGEL-00220/2020\|EPI_ISL_681968\|2020 | ···················· | ···················· | ···················· | ···················· | ···················· | ···················· | |
| U | 1516 hCoV-19/USA/PA-MGEL-00221/2020\|EPI_ISL_681969\|2020 | ···················· | ···················· | ···················· | ···················· | ···················· | ···················· | |
| U | 1517 hCoV-19/USA/PA-MGEL-00223/2020\|EPI_ISL_681970\|2020 | ···················· | ···················· | ···················· | ···················· | ···················· | ···················· | |
| U | 1518 hCoV-19/USA/PA-MGEL-00224/2020\|EPI_ISL_681971\|2020 | ···················· | ···················· | ···················· | ···················· | ···················· | ···················· | |
| U | 1519 hCoV-19/USA/PA-MGEL-00226/2020\|EPI_ISL_681972\|2020 | ···················· | ···················· | ···················· | ···················· | ···················· | ···················· | |
| U | 1520 hCoV-19/USA/PA-MGEL-00227/2020\|EPI_ISL_681973\|2020 | ···················· | ···················· | ···················· | ···················· | ···················· | ···················· | |
| U | 1521 hCoV-19/USA/PA-MGEL-00228/2020\|EPI_ISL_681974\|2020 | ···················· | ···················· | ···················· | ···················· | ···················· | ···················· | |
| U | 1522 hCoV-19/USA/PA-MGEL-00229/2020\|EPI_ISL_681975\|2020 | ···················· | ···················· | ···················· | ···················· | ···················· | ···················· | |
| U | 1523 hCoV-19/USA/PA-MGEL-00230/2020\|EPI_ISL_681976\|2020 | ···················· | ···················· | ···················· | ···················· | ···················· | ···················· | |
| U | 1524 hCoV-19/USA/PA-MGEL-00232/2020\|EPI_ISL_681977\|2020 | ···················· | ···················· | ···················· | ···················· | ···················· | ···················· | |
| U | 1525 hCoV-19/USA/PA-MGEL-00237/2020\|EPI_ISL_681978\|2020 | ···················· | ···················· | ···················· | ···················· | ···················· | ···················· | |
| U | 1526 hCoV-19/USA/PA-MGEL-00239/2020\|EPI_ISL_681979\|2020 | ···················· | ···················· | ···················· | ···················· | ···················· | ···················· | |
| U | 1527 hCoV-19/USA/PA-MGEL-00240/2020\|EPI_ISL_681980\|2020 | ···················· | ···················· | ···················· | ···················· | ···················· | ···················· | |
| U | 1528 hCoV-19/USA/PA-MGEL-00242/2020\|EPI_ISL_681981\|2020 | ···················· | ···················· | ···················· | ···················· | ···················· | ···················· | |
| U | 1529 hCoV-19/USA/PA-MGEL-00245/2020\|EPI_ISL_681982\|2020 | ···················· | ···················· | ···················· | ···················· | ···················· | ···················· | |
| U | 1530 hCoV-19/USA/PA-MGEL-00246/2020\|EPI_ISL_681983\|2020 | ···················· | ···················· | ···················· | ···················· | ···················· | ···················· | |
| U | 1531 hCoV-19/USA/PA-MGEL-00248/2020\|EPI_ISL_681984\|2020 | ···················· | ···················· | ···················· | ···················· | ···················· | ···················· | |
| U | 1532 hCoV-19/USA/PA-MGEL-00252/2020\|EPI_ISL_681985\|2020 | ···················· | ···················· | ···················· | ···················· | ···················· | ···················· | |
| U | 1533 hCoV-19/USA/PA-MGEL-00253/2020\|EPI_ISL_681986\|2020 | ···················· | ···················· | ···················· | ···················· | ···················· | ···················· | |
| U | 1534 hCoV-19/USA/PA-MGEL-00254/2020\|EPI_ISL_681987\|2020 | ···················· | ···················· | ···················· | ···················· | ···················· | ···················· | |
| U | 1535 hCoV-19/USA/PA-MGEL-00255/2020\|EPI_ISL_681988\|2020 | ···················· | ···················· | ···················· | ···················· | ···················· | ···················· | |
| U | 1536 hCoV-19/USA/PA-MGEL-00256/2020\|EPI_ISL_681989\|2020 | ···················· | ···················· | ···················· | ···················· | ···················· | ···················· | |
| U | 1537 hCoV-19/England/CAMC-B9AE24/2020\|EPI_ISL_673389\|20 | ···················· | ···················· | ···················· | ···················· | ···················· | ···················· | |
| U | 1538 hCoV-19/England/CAMC-B9A98D/2020\|EPI_ISL_673390\|20 | ···················· | ···················· | ···················· | ···················· | ···················· | ···················· | |
| U | 1539 hCoV-19/England/CAMC-B9A9BA/2020\|EPI_ISL_673391\|20 | ···················· | ···················· | ···················· | ···················· | ···················· | ···················· | |
| U | 1540 hCoV-19/England/CAMC-B9A2BF/2020\|EPI_ISL_673394\|20 | ···················· | ···················· | ···················· | ···················· | ···················· | ···················· | |
| U | 1541 hCoV-19/England/CAMC-B99098/2020\|EPI_ISL_673395\|20 | ···················· | ···················· | ···················· | ···················· | ···················· | ···················· | |
| U | 1542 hCoV-19/England/CAMC-B9C62A/2020\|EPI_ISL_673397\|20 | ···················· | ···················· | ···················· | ···················· | ···················· | ···················· | |

U 1543 hCoV-19/England/CAMC-B99520/2020|EPI_ISL_673398|20 ···················· ···················· ····················· ···················· ···················· ····················
U 1544 hCoV-19/England/CAMC-B98F14/2020|EPI_ISL_673422|20 ···················· ···················· ····················· ···················· ···················· ····················
U 1545 hCoV-19/England/CAMC-B9958A/2020|EPI_ISL_673423|20 ···················· ···················· ····················· ···················· ···················· ····················
U 1546 hCoV-19/England/CAMC-B9C4D5/2020|EPI_ISL_673424|20 ···················· ···················· ····················· ···················· ···················· ····················
U 1547 hCoV-19/England/CAMC-B9ADCD/2020|EPI_ISL_673427|20 ···················· ···················· ····················· ···················· ······T············· ····················
U 1548 hCoV-19/England/CAMC-B9B01E/2020|EPI_ISL_673428|20 ···················· ···················· ····················· ···················· ···················· ····················
U 1549 hCoV-19/England/CAMC-B992CF/2020|EPI_ISL_673429|20 ···················· ···················· ····················· ···················· ···················· ····················
U 1550 hCoV-19/England/CAMC-B9A03D/2020|EPI_ISL_673430|20 ···················· ···················· ····················· ···················· ······T············· ····················
U 1551 hCoV-19/England/CAMC-B9A923/2020|EPI_ISL_673431|20 ···················· ···················· ····················· ···················· ···················· ····················
U 1552 hCoV-19/England/CAMC-B98EEA/2020|EPI_ISL_673433|20 ···················· ···················· ····················· ···················· ···················· ····················
U 1553 hCoV-19/England/CAMC-B99D55/2020|EPI_ISL_673440|20 ···················· ···················· ····················· ···················· ···················· ····················
U 1554 hCoV-19/England/CAMC-B9AE06/2020|EPI_ISL_673441|20 ···················· ···················· ····················· ···················· ···················· ····················
U 1555 hCoV-19/England/CAMC-B99863/2020|EPI_ISL_673442|20 ···················· ···················· ····················· ···················· ···················· ····················
U 1556 hCoV-19/England/MILK-B8F7EB/2020|EPI_ISL_674753|20 ···················· ···················· ····················· ···················· ···················· ····················
U 1557 hCoV-19/England/MILK-B8EE05/2020|EPI_ISL_674754|20 ···················· ···················· ····················· ···················· ···················· ····················
U 1558 hCoV-19/England/MILK-B8EC83/2020|EPI_ISL_674755|20 ···················· ···················· ····················· ···················· ···················· ····················
U 1559 hCoV-19/England/QEUH-B7CEEC/2020|EPI_ISL_674756|20 ···················· ···················· ····················· ···················· ···················· ····················
U 1560 hCoV-19/England/MILK-B8EB1D/2020|EPI_ISL_674757|20 ···················· ···················· ····················· ···················· ···················· ····················
U 1561 hCoV-19/England/MILK-B8E9D7/2020|EPI_ISL_674761|20 ···················· ···················· ····················· ···················· ···················· ····················
U 1562 hCoV-19/England/QEUH-B8089C/2020|EPI_ISL_674762|20 ···················· ···················· ····················· ···················· ···················· ····················
U 1563 hCoV-19/England/QEUH-B7F872/2020|EPI_ISL_674763|20 ···················· ···················· ····················· ···················· ···················· ····················
U 1564 hCoV-19/England/MILK-B8E03C/2020|EPI_ISL_674771|20 ···················· ···················· ····················· ···················· ···················· ····················
U 1565 hCoV-19/England/QEUH-B7F766/2020|EPI_ISL_674772|20 ···················· ···················· ····················· ···················· ···················· ····················
U 1566 hCoV-19/England/QEUH-B808AB/2020|EPI_ISL_674773|20 ···················· ·A·················· ····················· ···················· ···················· ····················
U 1567 hCoV-19/England/MILK-B8E4E5/2020|EPI_ISL_674774|20 ···················· ···················· ····················· ···················· ···················· ····················
U 1568 hCoV-19/Saudi Arabia/KAUST-RIYADH1194/2020|EPI_ISL ···················· ···················· ····················· ···················· ···················· ····················
U 1569 hCoV-19/Saudi Arabia/KAUST-RIYADH1195/2020|EPI_ISL ···················· ·A·················· ····················· ···················· ···················· ····················
U 1570 hCoV-19/Saudi Arabia/KAUST-RIYADH1196/2020|EPI_ISL ···················· ···················· ····················· ···················· ···················· ···A················
U 1571 hCoV-19/Saudi Arabia/KAUST-RIYADH1205/2020|EPI_ISL ···················· ···················· ····················· ···················· ···················· ····················
U 1572 hCoV-19/Saudi Arabia/KAUST-RIYADH1207/2020|EPI_ISL ···················· ···················· ····················· ···················· ···················· ····················
U 1573 hCoV-19/Saudi Arabia/KAUST-RIYADH1208/2020|EPI_ISL ···················· ···················· ····················· ···················· ···················· ····················
U 1574 hCoV-19/Saudi Arabia/KAUST-RIYADH1214/2020|EPI_ISL ···················· ·A·················· ····················· ···················· ···················· ····················
U 1575 hCoV-19/Saudi Arabia/KAUST-RIYADH1215/2020|EPI_ISL ···················· ·A·················· ····················· ···················· ···················· ···A················
U 1576 hCoV-19/Saudi Arabia/KAUST-RIYADH1216/2020|EPI_ISL ···················· ···················· ····················· ···················· ···················· ····················
U 1577 hCoV-19/Saudi Arabia/KAUST-RIYADH1219/2020|EPI_ISL ···················· ···················· ····················· ···················· ···················· ····················
U 1578 hCoV-19/Saudi Arabia/KAUST-RIYADH1220/2020|EPI_ISL ···················· ···················· ····················· ···················· ···················· ····················
U 1579 hCoV-19/Saudi Arabia/KAUST-RIYADH1222/2020|EPI_ISL ···················· ·A·················· ····················· ···················· ···················· ····················
U 1580 hCoV-19/Saudi Arabia/KAUST-RIYADH1235/2020|EPI_ISL ···················· ·A·················· ····················· ···················· ···················· ····················
U 1581 hCoV-19/Saudi Arabia/KAUST-RIYADH1241/2020|EPI_ISL ···················· ·A·················· ····················· ···················· ···················· ···A················
U 1582 hCoV-19/Saudi Arabia/KAUST-RIYADH1243/2020|EPI_ISL ···················· ·A·················· ····················· ···················· ···················· ····················
U 1583 hCoV-19/Saudi Arabia/KAUST-RIYADH1249/2020|EPI_ISL ···················· ···················· ····················· ···················· ···················· ····················
U 1584 hCoV-19/Saudi Arabia/KAUST-RIYADH1250/2020|EPI_ISL ···················· ···················· ····················· ···················· ···················· ····················
U 1585 hCoV-19/Saudi Arabia/KAUST-RIYADH1251/2020|EPI_ISL ···················· ·A·················· ····················· ···················· ···················· ····················
U 1586 hCoV-19/Saudi Arabia/KAUST-RIYADH1254/2020|EPI_ISL ···················· ·A·················· ····················· ···················· ···················· ···A················
U 1587 hCoV-19/Saudi Arabia/KAUST-RIYADH1259/2020|EPI_ISL ···················· ·A·················· ····················· ···················· ···················· ···A················
U 1588 hCoV-19/Saudi Arabia/KAUST-RIYADH1261/2020|EPI_ISL ···················· ···················· ····················· ···················· ···················· ····················
U 1589 hCoV-19/Saudi Arabia/KAUST-RIYADH1263/2020|EPI_ISL ···················· ·A·················· ····················· ···················· ···················· ····················
U 1590 hCoV-19/Saudi Arabia/KAUST-RIYADH1264/2020|EPI_ISL ···················· ···················· ····················· ···················· ···················· ····················
U 1591 hCoV-19/Saudi Arabia/KAUST-RIYADH1265/2020|EPI_ISL ···················· ·A·················· ····················· ···················· ···················· ···A················
U 1592 hCoV-19/Saudi Arabia/KAUST-RIYADH1267/2020|EPI_ISL ···················· ·A·················· ····················· ···················· ···················· ···A················
U 1593 hCoV-19/Saudi Arabia/KAUST-RIYADH1268/2020|EPI_ISL ···················· ·A·················· ····················· ···················· ···················· ···A················
U 1594 hCoV-19/Saudi Arabia/KAUST-RIYADH1442/2020|EPI_ISL ···················· ···················· ····················· ···················· ···················· ····················
U 1596 hCoV-19/Saudi Arabia/KAUST-MAKKAH185/2020|EPI_ISL_ ···················· ···················· ····················· ···················· ···················· ····················
U 1598 hCoV-19/Saudi Arabia/KAUST-MADINAH1132/2020|EPI_IS ···················· ·A·················· ····················· ···················· ···················· ···A················
U 1599 hCoV-19/Saudi Arabia/KAUST-RIYADH1240/2020|EPI_ISL ···················· ·A·················· ····················· ···················· ···················· ····················
U 1600 hCoV-19/Saudi Arabia/KAUST-JEDDAH1293/2020|EPI_ISL ···················· ·A·················· ····················· ···················· ···················· ···A················
U 1601 hCoV-19/Saudi Arabia/KAUST-MAKKAH1610/2020|EPI_ISL ···················· ···················· ····················· ···················· ···················· ····················
U 1602 hCoV-19/Saudi Arabia/KAUST-RIYADH1233/2020|EPI_ISL ···················· ·A·················· ····················· ···················· ···················· ···A················
U 1603 hCoV-19/North Macedonia/9451/2020|EPI_ISL_678248|2 ···················· ···················· ····················· ···················· ···················· ····················
U 1604 hCoV-19/North Macedonia/9457/2020|EPI_ISL_678249|2 ···················· ···················· ····················· ···················· ···················· ····················
U 1605 hCoV-19/North Macedonia/9675/2020|EPI_ISL_678250|2 ···················· ···················· ····················· ···················· ···················· ····················
U 1606 hCoV-19/North Macedonia/10299/2020|EPI_ISL_678251| ···················· ···················· ····················· ···················· ···················· ····················
U 1607 hCoV-19/North Macedonia/10328/2020|EPI_ISL_678252| ···················· ···················· ····················· ···················· ···················· ····················
U 1608 hCoV-19/North Macedonia/11567/2020|EPI_ISL_678253| ···················· ···················· ····················· ···················· ···················· ····················
U 1609 hCoV-19/North Macedonia/11602/2020|EPI_ISL_678254| ···················· ···················· ····················· ···················· ···················· ····················
U 1610 hCoV-19/North Macedonia/11633/2020|EPI_ISL_678255| ···················· ···················· ····················· ···················· ···················· ····················
U 1611 hCoV-19/North Macedonia/11638/2020|EPI_ISL_678256| ···················· ···················· ····················· ···················· ·········T·········· ····················
U 1612 hCoV-19/North Macedonia/11777/2020|EPI_ISL_678257| ···················· ···················· ····················· ···················· ···················· ····················
U 1613 hCoV-19/North Macedonia/11805/2020|EPI_ISL_678258| ···················· ···················· ····················· ···················· ···················· ····················
U 1614 hCoV-19/North Macedonia/11890/2020|EPI_ISL_678259| ···················· ···················· ····················· ···················· ···················· ····················
U 1615 hCoV-19/North Macedonia/12026/2020|EPI_ISL_678260| ···················· ···················· ····················· ···················· ···················· ····················
U 1616 hCoV-19/Bahrain/600041988_S21_L001/2020|EPI_ISL_67 ···················· ·A·················· ····················· ···················· ···················· ···A················
U 1617 hCoV-19/Bahrain/250182770_S11_L001/2020|EPI_ISL_67 ···················· ···················· ····················· ···················· ···················· ····················
U 1618 hCoV-19/Bahrain/341013928_S4_L001/2020|EPI_ISL_678 ···················· ···················· ····················· ···················· ···················· ····················
U 1619 hCoV-19/Bahrain/341016741_S2_L001/2020|EPI_ISL_678 ···················· ···················· ····················· ···················· ···················· ····················
U 1620 hCoV-19/Bahrain/341016804_S1_L001/2020|EPI_ISL_678 ···················· ···················· ····················· ···················· ···················· ····················
U 1621 hCoV-19/Bahrain/341038078_S3_L001/2020|EPI_ISL_678 ···················· ···················· ····················· ···················· ···················· ····················
U 1622 hCoV-19/Bahrain/341051639_S14_L001/2020|EPI_ISL_67 ···················· ···················· ····················· ···················· ···················· ····················
U 1623 hCoV-19/Bahrain/341051813_S12_L001/2020|EPI_ISL_67 ···················· ···················· ····················· ···················· ···················· ····················
U 1624 hCoV-19/Bahrain/341063114_S18_L001/2020|EPI_ISL_67 ···················· ···················· ····················· ···················· ···················· ····················

| | | | | | | | | |
|---|---|---|---|---|---|---|---|---|
| U | 1625 | hCoV-19/Bahrain/341063655_S20_L001/2020\|EPI_ISL_67 | ···················· | ···A················ | ···················· | ···················· | ···················· | ··A················· |
| U | 1626 | hCoV-19/Bahrain/341063863_S17_L001/2020\|EPI_ISL_67 | ···················· | ···················· | ···················· | ···················· | ···················· | ···················· |
| U | 1627 | hCoV-19/Bahrain/341063898_S19_L001/2020\|EPI_ISL_67 | ···················· | ···················· | ···················· | ···················· | ···················· | ···················· |
| U | 1628 | hCoV-19/Russia/SAR-RARI-1150/2020\|EPI_ISL_678273\|2 | ···················· | ···················· | ···················· | ···················· | ···················· | ···················· |
| U | 1629 | hCoV-19/Russia/SAR-RARI-3187/2020\|EPI_ISL_678274\|2 | ···················· | ···················· | ···················· | ···················· | ···················· | ···················· |
| U | 1630 | hCoV-19/Russia/MOS-RARI-1054/2020\|EPI_ISL_678275\|2 | ···················· | ···················· | ···················· | ···················· | ···················· | ···················· |
| U | 1631 | hCoV-19/Russia/SAM-RARI-1011/2020\|EPI_ISL_678276\|2 | ···················· | ···················· | ···················· | ···················· | ···················· | ···················· |
| U | 1632 | hCoV-19/Russia/SAR-RARI-3663/2020\|EPI_ISL_678277\|2 | ···················· | ···················· | ···················· | ···················· | ···················· | ···················· |
| U | 1633 | hCoV-19/Russia/MOS-RARI-3709/2020\|EPI_ISL_678278\|2 | ···················· | ···················· | ···················· | ···················· | ···················· | ···················· |
| U | 1634 | hCoV-19/Russia/MOS-RARI-879/2020\|EPI_ISL_678279\|20 | ···················· | ···················· | ···················· | ···················· | ···················· | ···················· |
| U | 1635 | hCoV-19/England/LOND-1263EFD/2020\|EPI_ISL_679502\|2 | ···················· | ·A·················· | ···················· | ···················· | ···················· | ··A················· |
| U | 1636 | hCoV-19/England/LOND-1263F90/2020\|EPI_ISL_679508\|2 | ···················· | ···················· | ···················· | ···················· | ···················· | ···················· |
| U | 1637 | hCoV-19/England/LOND-1263FDC/2020\|EPI_ISL_679511\|2 | ···················· | ···················· | ···················· | ···················· | ···················· | ···················· |
| U | 1638 | hCoV-19/England/LOND-1264032/2020\|EPI_ISL_679514\|2 | ···················· | ···················· | ···················· | ···················· | ·····T·············· | ···················· |
| U | 1639 | hCoV-19/England/LOND-1264041/2020\|EPI_ISL_679515\|2 | ···················· | ···················· | ···················· | ···················· | ···················· | ···················· |
| U | 1640 | hCoV-19/England/LOND-126407E/2020\|EPI_ISL_679518\|2 | ···················· | ·A·················· | ···················· | ···················· | ···················· | ··A················· |
| U | 1641 | hCoV-19/England/LOND-1264111/2020\|EPI_ISL_679526\|2 | ···················· | ·A·················· | ···················· | ···················· | ···················· | ··A················· |
| U | 1642 | hCoV-19/England/LOND-12D4BA5/2020\|EPI_ISL_679551\|2 | ···················· | ···················· | ···················· | ···················· | ···················· | ···················· |
| U | 1643 | hCoV-19/England/LOND-12D4BE1/2020\|EPI_ISL_679555\|2 | ···················· | ···················· | ···················· | ···················· | ···················· | ···················· |
| U | 1644 | hCoV-19/England/LOND-12D4C0C/2020\|EPI_ISL_679557\|2 | ···················· | ···················· | ···················· | ···················· | ···················· | ···················· |
| U | 1645 | hCoV-19/Scotland/EDB10199/2020\|EPI_ISL_680500\|2020 | ···················· | ···················· | ···················· | ···················· | ···················· | ···················· |
| U | 1646 | hCoV-19/Scotland/EDB10201/2020\|EPI_ISL_680502\|2020 | ···················· | ···················· | ···················· | ···················· | ···················· | ···················· |
| U | 1647 | hCoV-19/Scotland/EDB10205/2020\|EPI_ISL_680506\|2020 | ···················· | ···················· | ···················· | ···················· | ···················· | ···················· |
| U | 1648 | hCoV-19/Scotland/EDB10206/2020\|EPI_ISL_680507\|2020 | ···················· | ···················· | ···················· | ···················· | ···················· | ···················· |
| U | 1650 | hCoV-19/Scotland/EDB10209/2020\|EPI_ISL_680509\|2020 | ···················· | ···················· | ···················· | ···················· | ···················· | ···················· |
| U | 1652 | hCoV-19/Scotland/EDB9082/2020\|EPI_ISL_680511\|2020- | ···················· | ···················· | ···················· | ···················· | ···················· | ···················· |
| U | 1653 | hCoV-19/Scotland/EDB9402/2020\|EPI_ISL_680513\|2020- | ···················· | ···················· | ···················· | ···················· | ·····T·············· | ···················· |
| U | 1654 | hCoV-19/Scotland/EDB9403/2020\|EPI_ISL_680514\|2020- | ···················· | ···················· | ···················· | ···················· | ···················· | ···················· |
| U | 1655 | hCoV-19/Scotland/EDB9408/2020\|EPI_ISL_680515\|2020- | ···················· | ···················· | ···················· | ···················· | ···················· | ···················· |
| U | 1656 | hCoV-19/Scotland/EDB9410/2020\|EPI_ISL_680516\|2020- | ···················· | ···················· | ···················· | ···················· | ·····T·············· | ···················· |
| U | 1657 | hCoV-19/Scotland/EDB9412/2020\|EPI_ISL_680517\|2020- | ···················· | ···················· | ···················· | ···················· | ·····T·············· | ···················· |
| U | 1658 | hCoV-19/Scotland/EDB9414/2020\|EPI_ISL_680518\|2020- | ···················· | ···················· | ···················· | ···················· | ·····T·············· | ···················· |
| U | 1659 | hCoV-19/Scotland/EDB9415/2020\|EPI_ISL_680519\|2020- | ···················· | ···················· | ···················· | ···················· | ·····T·············· | ···················· |
| U | 1660 | hCoV-19/Scotland/EDB9417/2020\|EPI_ISL_680521\|2020- | ···················· | ···················· | ···················· | ···················· | ·····T·············· | ···················· |
| U | 1661 | hCoV-19/Scotland/EDB9419/2020\|EPI_ISL_680522\|2020- | ···················· | ···················· | ···················· | ···················· | ·····T·············· | ···················· |
| U | 1662 | hCoV-19/Scotland/EDB9430/2020\|EPI_ISL_680524\|2020- | ···················· | ···················· | ···················· | ···················· | ·····T·············· | ···················· |
| U | 1663 | hCoV-19/Scotland/EDB9431/2020\|EPI_ISL_680525\|2020- | ···················· | ···················· | ···················· | ···················· | ·····T·············· | ···················· |
| U | 1664 | hCoV-19/Scotland/EDB9433/2020\|EPI_ISL_680526\|2020- | ···················· | ···················· | ···················· | ···················· | ·····T·············· | ···················· |
| U | 1665 | hCoV-19/Scotland/EDB9434/2020\|EPI_ISL_680527\|2020- | ···················· | ···················· | ···················· | ···················· | ·····T·············· | ···················· |
| U | 1666 | hCoV-19/Scotland/EDB9435/2020\|EPI_ISL_680528\|2020- | ···················· | ···················· | ···················· | ···················· | ·····T·············· | ···················· |
| U | 1667 | hCoV-19/Scotland/EDB9438/2020\|EPI_ISL_680529\|2020- | ···················· | ···················· | ···················· | ···················· | ·····T·············· | ···················· |
| U | 1668 | hCoV-19/Scotland/EDB9439/2020\|EPI_ISL_680530\|2020- | ···················· | ···················· | ···················· | ···················· | ·····T·············· | ···················· |
| U | 1669 | hCoV-19/Scotland/EDB9442/2020\|EPI_ISL_680531\|2020- | ···················· | ···················· | ···················· | ···················· | ·····T·············· | ···················· |
| U | 1670 | hCoV-19/Scotland/EDB9443/2020\|EPI_ISL_680532\|2020- | ···················· | ···················· | ···················· | ···················· | ·····T·············· | ···················· |
| U | 1671 | hCoV-19/Scotland/EDB9445/2020\|EPI_ISL_680533\|2020- | ···················· | ···················· | ···················· | ···················· | ·····T·············· | ···················· |
| U | 1673 | hCoV-19/Scotland/EDB9452/2020\|EPI_ISL_680536\|2020- | ···················· | ···················· | ···················· | ···················· | ···················· | ···················· |
| U | 1674 | hCoV-19/Scotland/EDB9463/2020\|EPI_ISL_680537\|2020- | ···················· | ···················· | ···················· | ···················· | ·····T·············· | ···················· |
| U | 1675 | hCoV-19/Scotland/EDB9671/2020\|EPI_ISL_680538\|2020- | ···················· | ·A·······A·········· | ···················· | ···················· | ···················· | ···················· |
| U | 1676 | hCoV-19/Scotland/EDB9746/2020\|EPI_ISL_680539\|2020- | ···················· | ···················· | ···················· | ···················· | ···················· | ···················· |
| U | 1678 | hCoV-19/Scotland/EDB9798/2020\|EPI_ISL_680541\|2020- | ···················· | ···················· | ···················· | ···················· | ···················· | ···················· |
| U | 1679 | hCoV-19/Scotland/EDB9803/2020\|EPI_ISL_680542\|2020- | ···················· | ···················· | ···················· | ···················· | ···················· | ···················· |
| U | 1681 | hCoV-19/Scotland/EDB9874/2020\|EPI_ISL_680546\|2020- | ···················· | ···················· | ···················· | ···················· | ···················· | ···················· |
| U | 1682 | hCoV-19/Scotland/EDB9875/2020\|EPI_ISL_680547\|2020- | ···················· | ···················· | ···················· | ···················· | ·····T·············· | ···················· |
| U | 1683 | hCoV-19/Scotland/EDB9930/2020\|EPI_ISL_680549\|2020- | ···················· | ···················· | ···················· | ···················· | ···················· | ···················· |
| U | 1684 | hCoV-19/Scotland/EDB9942/2020\|EPI_ISL_680550\|2020- | ···················· | ···················· | ···················· | ···················· | ···················· | ···················· |
| U | 1685 | hCoV-19/Scotland/EDB9949/2020\|EPI_ISL_680551\|2020- | ···················· | ···················· | ···················· | ···················· | ···················· | ···················· |
| U | 1686 | hCoV-19/Scotland/EDB9951/2020\|EPI_ISL_680552\|2020- | ···················· | ···················· | ···················· | ···················· | ···················· | ···················· |
| U | 1687 | hCoV-19/Scotland/EDB9964/2020\|EPI_ISL_680553\|2020- | ···················· | ···················· | ···················· | ···················· | ···················· | ···················· |
| U | 1688 | hCoV-19/Scotland/EDB9972/2020\|EPI_ISL_680554\|2020- | ···················· | ···················· | ···················· | ···················· | ···················· | ···················· |
| U | 1689 | hCoV-19/Scotland/EDB9979/2020\|EPI_ISL_680555\|2020- | ···················· | ···················· | ···················· | ···················· | ···················· | ···················· |
| U | 1690 | hCoV-19/Scotland/QEUH-A016D6/2020\|EPI_ISL_680561\|2 | ···················· | ···················· | ···················· | ···················· | ···················· | ···················· |
| U | 1691 | hCoV-19/Scotland/QEUH-A9D2EA/2020\|EPI_ISL_680564\|2 | ···················· | ···················· | ···················· | ···················· | ···················· | ···················· |
| U | 1692 | hCoV-19/Scotland/QEUH-B2C23A/2020\|EPI_ISL_680571\|2 | ···················· | ···················· | ···················· | ···················· | ···················· | ···················· |
| U | 1693 | hCoV-19/Scotland/QEUH-B2C294/2020\|EPI_ISL_680572\|2 | ···················· | ···················· | ···················· | ···················· | ···················· | ···················· |
| U | 1694 | hCoV-19/Scotland/QEUH-B2C2C1/2020\|EPI_ISL_680573\|2 | ···················· | ···················· | ···················· | ···················· | ···················· | ···················· |
| U | 1695 | hCoV-19/Scotland/QEUH-B2CF33/2020\|EPI_ISL_680574\|2 | ···················· | ···················· | ···················· | ···················· | ···················· | ···················· |
| U | 1696 | hCoV-19/Scotland/QEUH-B2D020/2020\|EPI_ISL_680576\|2 | ···················· | ···················· | ···················· | ···················· | ···················· | ···················· |
| U | 1697 | hCoV-19/Wales/MILK-B3A8DD/2020\|EPI_ISL_680579\|2020 | ···················· | ···················· | ···················· | ···················· | ···················· | ···················· |
| U | 1700 | hCoV-19/Wales/PHWC-4880B1/2020\|EPI_ISL_680582\|2020 | ···················· | ···················· | ···················· | ···················· | ···················· | ···················· |
| U | 1701 | hCoV-19/Wales/PHWC-48B995/2020\|EPI_ISL_680584\|2020 | ···················· | ···················· | ···················· | ···················· | ···················· | ···················· |
| U | 1702 | hCoV-19/Wales/PHWC-48BA65/2020\|EPI_ISL_680585\|2020 | ···················· | ···················· | ···················· | ···················· | ···················· | ···················· |
| U | 1703 | hCoV-19/Wales/PHWC-48C712/2020\|EPI_ISL_680586\|2020 | ···················· | ···················· | ···················· | ···················· | ···················· | ···················· |
| U | 1704 | hCoV-19/Wales/PHWC-48D1C8/2020\|EPI_ISL_680587\|2020 | ···················· | ···················· | ···················· | ···················· | ·····T·············· | ···················· |
| U | 1705 | hCoV-19/Wales/PHWC-48D1D7/2020\|EPI_ISL_680588\|2020 | ···················· | ···················· | ···················· | ···················· | ···················· | ···················· |
| U | 1706 | hCoV-19/Wales/PHWC-48D1F5/2020\|EPI_ISL_680590\|2020 | ···················· | ···················· | ···················· | ···················· | ···················· | ···················· |
| U | 1707 | hCoV-19/Wales/PHWC-48D201/2020\|EPI_ISL_680591\|2020 | ···················· | ···················· | ···················· | ···················· | ···················· | ···················· |
| U | 1708 | hCoV-19/Wales/PHWC-48D22F/2020\|EPI_ISL_680593\|2020 | ···················· | ···················· | ···················· | ···················· | ···················· | ···················· |
| U | 1709 | hCoV-19/USA/GA-CDC-7669/2020\|EPI_ISL_681800\|2020-1 | ···················· | ···················· | ···················· | ···················· | ···················· | ···················· |
| U | 1710 | hCoV-19/USA/GA-CDC-7671/2020\|EPI_ISL_681801\|2020-1 | ···················· | ···················· | ···················· | ···················· | ···················· | ···················· |
| U | 1711 | hCoV-19/USA/GA-CDC-7701/2020\|EPI_ISL_681802\|2020-1 | ···················· | ···················· | ···················· | ···················· | ···················· | ···················· |

| | | | | | | | | |
|---|---|---|---|---|---|---|---|---|
| U | 1712 | hCoV-19/USA/GA-CDC-7704/2020\|EPI_ISL_681803\|2020-1 | .................... | .................... | .................... | .................... | .................... | .................... |
| U | 1713 | hCoV-19/USA/GA-CDC-7711/2020\|EPI_ISL_681804\|2020-1 | .................... | .................... | .................... | .................... | .................... | .................... |
| U | 1714 | hCoV-19/USA/GA-CDC-7720/2020\|EPI_ISL_681805\|2020-1 | .................... | .................... | .................... | .................... | .................... | .................... |
| U | 1715 | hCoV-19/USA/GA-CDC-7724/2020\|EPI_ISL_681806\|2020-1 | .................... | .................... | .................... | .................... | .................... | .................... |
| U | 1716 | hCoV-19/USA/GA-CDC-7728/2020\|EPI_ISL_681807\|2020-1 | .................... | .................... | .................... | .................... | .................... | .................... |
| U | 1717 | hCoV-19/USA/GA-CDC-7731/2020\|EPI_ISL_681808\|2020-1 | .................... | .................... | .................... | .................... | .................... | .................... |
| U | 1718 | hCoV-19/USA/GA-CDC-7733/2020\|EPI_ISL_681809\|2020-1 | .................... | .................... | .................... | .................... | .................... | .................... |
| U | 1719 | hCoV-19/USA/GA-CDC-7742/2020\|EPI_ISL_681810\|2020-1 | .................... | .................... | .................... | .................... | .................... | .................... |
| U | 1720 | hCoV-19/USA/GA-CDC-7743/2020\|EPI_ISL_681811\|2020-1 | .................... | .................... | .................... | .................... | .................... | .................... |
| U | 1721 | hCoV-19/USA/GA-CDC-7744/2020\|EPI_ISL_681812\|2020-1 | .................... | .................... | .................... | .................... | .................... | .................... |
| U | 1722 | hCoV-19/USA/GA-CDC-7746/2020\|EPI_ISL_681813\|2020-1 | .................... | .................... | .................... | .................... | .................... | .................... |
| U | 1723 | hCoV-19/USA/GA-CDC-7799/2020\|EPI_ISL_681814\|2020-1 | .................... | .................... | .................... | .................... | .................... | .................... |
| U | 1724 | hCoV-19/USA/GA-CDC-7823/2020\|EPI_ISL_681815\|2020-1 | .................... | .................... | .................... | .................... | .................... | .................... |
| U | 1725 | hCoV-19/USA/GA-CDC-7877/2020\|EPI_ISL_681816\|2020-1 | .................... | .................... | .................... | .................... | .................... | .................... |
| U | 1726 | hCoV-19/USA/GA-CDC-7896/2020\|EPI_ISL_681817\|2020-1 | .................... | .................... | .................... | .................... | .................... | .................... |
| U | 1727 | hCoV-19/USA/GA-CDC-8142/2020\|EPI_ISL_681818\|2020-1 | .................... | .................... | .................... | .................... | .................... | .................... |
| U | 1728 | hCoV-19/USA/GA-CDC-7402/2020\|EPI_ISL_681819\|2020-1 | .................... | .................... | .................... | .................... | .................... | .................... |
| U | 1729 | hCoV-19/USA/GA-CDC-7407/2020\|EPI_ISL_681820\|2020-1 | .................... | .................... | .................... | .................... | .................... | .................... |
| U | 1730 | hCoV-19/USA/GA-CDC-7459/2020\|EPI_ISL_681821\|2020-1 | .................... | .................... | .................... | .................... | .................... | .................... |
| U | 1731 | hCoV-19/USA/GA-CDC-7675/2020\|EPI_ISL_681822\|2020-1 | .................... | .................... | .................... | .................... | .................... | .................... |
| U | 1732 | hCoV-19/USA/GA-CDC-7676/2020\|EPI_ISL_681823\|2020-1 | .................... | .................... | .................... | .................... | .................... | .................... |
| U | 1733 | hCoV-19/USA/GA-CDC-7679/2020\|EPI_ISL_681824\|2020-1 | .................... | .................... | .................... | .................... | .................... | .................... |
| U | 1734 | hCoV-19/USA/GA-CDC-7681/2020\|EPI_ISL_681825\|2020-1 | .................... | .................... | .................... | .................... | .................... | .................... |
| U | 1735 | hCoV-19/USA/GA-CDC-7845/2020\|EPI_ISL_681826\|2020-1 | .................... | .................... | .................... | .................... | .................... | .................... |
| U | 1736 | hCoV-19/USA/GA-CDC-7860/2020\|EPI_ISL_681827\|2020-1 | .................... | .................... | .................... | .................... | .................... | .................... |
| U | 1737 | hCoV-19/USA/GA-CDC-7867/2020\|EPI_ISL_681828\|2020-1 | .................... | .................... | .................... | .................... | .................... | .................... |
| U | 1738 | hCoV-19/Cote dIvoire/C01-2176/2020\|EPI_ISL_681829\| | .................... | .................... | .................... | .................... | .................... | .................... |
| U | 1741 | hCoV-19/Cote dIvoire/BKE0366/2020\|EPI_ISL_681834\|2 | .................... | .................... | .................... | .................... | .................... | .................... |
| U | 1742 | hCoV-19/Cote dIvoire/BKE0382/2020\|EPI_ISL_681835\|2 | .................... | .................... | .................... | .................... | .................... | .................... |
| U | 1743 | hCoV-19/Cote dIvoire/BKE0430/2020\|EPI_ISL_681836\|2 | .................... | .................... | .................... | .................... | .................... | .................... |
| U | 1744 | hCoV-19/Cote dIvoire/BKE0820/2020\|EPI_ISL_681840\|2 | .................... | .................... | .................... | .................... | .................... | .................... |
| U | 1745 | hCoV-19/Cote dIvoire/BKE0837/2020\|EPI_ISL_681841\|2 | .................... | .................... | .................... | .................... | .................... | .................... |
| U | 1746 | hCoV-19/USA/TX-DSHS-1544/2020\|EPI_ISL_681876\|2020- | .................... | .................... | .................... | .................... | .................... | .................... |
| U | 1747 | hCoV-19/USA/TX-DSHS-1553/2020\|EPI_ISL_681877\|2020- | .................... | .................... | .................... | .................... | .................... | .................... |
| U | 1748 | hCoV-19/USA/TX-DSHS-1556/2020\|EPI_ISL_681878\|2020- | .................... | .................... | .................... | .................... | .................... | .................... |
| U | 1749 | hCoV-19/USA/TX-DSHS-1557/2020\|EPI_ISL_681879\|2020- | .................... | .A.................. | .................... | .................... | .................... | .................... |
| U | 1750 | hCoV-19/USA/TX-DSHS-1558/2020\|EPI_ISL_681880\|2020- | .................... | .A.................. | .................... | .................... | .................... | .................... |
| U | 1751 | hCoV-19/USA/TX-DSHS-1564/2020\|EPI_ISL_681881\|2020- | .................... | .................... | .................... | .................... | .................... | .................... |
| U | 1752 | hCoV-19/USA/TX-DSHS-1574/2020\|EPI_ISL_681882\|2020- | .................... | .A.................. | .................... | .................... | .................... | .................... |
| U | 1753 | hCoV-19/USA/TX-DSHS-1576/2020\|EPI_ISL_681883\|2020- | .................... | .................... | .................... | .................... | .................... | .................... |
| U | 1754 | hCoV-19/Ireland/LD-NVRL-20G49957/2020\|EPI_ISL_6818 | .................... | .................... | .................... | .................... | .................... | .................... |
| U | 1755 | hCoV-19/Ireland/MN-NVRL-20Z06662/2020\|EPI_ISL_6818 | .................... | .................... | .................... | .................... | .................... | .................... |
| U | 1756 | hCoV-19/Ireland/MN-NVRL-20Z06692/2020\|EPI_ISL_6818 | .................... | .................... | .................... | .................... | .................... | .................... |
| U | 1757 | hCoV-19/Ireland/MN-NVRL-20Z06201/2020\|EPI_ISL_6818 | .................... | .................... | .................... | .................... | .................... | .................... |
| U | 1758 | hCoV-19/Ireland/MN-NVRL-20Z06211/2020\|EPI_ISL_6818 | .................... | .................... | .................... | .................... | .................... | .................... |
| U | 1759 | hCoV-19/Ireland/LS-NVRL-20Z06375/2020\|EPI_ISL_6818 | .................... | .................... | .................... | .................... | .................... | .................... |
| U | 1760 | hCoV-19/Ireland/D-NVRL-20Z06394/2020\|EPI_ISL_68189 | .................... | .................... | .................... | .................... | .................... | .................... |
| U | 1761 | hCoV-19/Costa Rica/INC-0095/2020\|EPI_ISL_682237\|20 | .................... | .................... | .................... | .................... | .................... | .................... |
| U | 1762 | hCoV-19/Costa Rica/INC-0100/2020\|EPI_ISL_682241\|20 | .................... | .................... | .................... | .................... | .................... | .................... |
| U | 1763 | hCoV-19/Costa Rica/INC-0101/2020\|EPI_ISL_682242\|20 | .................... | .................... | .................... | .................... | .................... | .................... |
| U | 1764 | hCoV-19/Costa Rica/INC-0102/2020\|EPI_ISL_682243\|20 | .................... | .................... | .................... | .................... | .................... | .................... |
| U | 1765 | hCoV-19/Costa Rica/INC-0103/2020\|EPI_ISL_682244\|20 | .................... | .................... | .................... | .................... | .................... | .................... |
| U | 1766 | hCoV-19/Costa Rica/INC-0104/2020\|EPI_ISL_682245\|20 | .................... | .................... | .................... | .................... | .................... | .................... |
| U | 1767 | hCoV-19/Costa Rica/INC-0105/2020\|EPI_ISL_682246\|20 | .................... | .................... | .................... | .................... | .................... | .................... |
| U | 1768 | hCoV-19/Costa Rica/INC-0107/2020\|EPI_ISL_682247\|20 | .................... | .................... | .................... | .................... | .................... | .................... |
| U | 1769 | hCoV-19/Costa Rica/INC-0108/2020\|EPI_ISL_682248\|20 | .................... | .................... | .................... | .................... | .................... | .................... |
| U | 1770 | hCoV-19/Costa Rica/INC-0109/2020\|EPI_ISL_682249\|20 | .................... | .................... | .................... | .................... | .................... | .................... |
| U | 1771 | hCoV-19/Costa Rica/INC-0110/2020\|EPI_ISL_682250\|20 | .................... | .................... | .................... | .................... | .................... | .................... |
| U | 1772 | hCoV-19/Costa Rica/INC-0112/2020\|EPI_ISL_682252\|20 | .................... | .................... | .................... | .................... | .................... | .................... |
| U | 1773 | hCoV-19/Costa Rica/INC-0113/2020\|EPI_ISL_682253\|20 | .................... | .................... | .................... | .................... | .................... | .................... |
| U | 1774 | hCoV-19/Costa Rica/INC-0114/2020\|EPI_ISL_682254\|20 | .................... | .................... | .................... | .................... | .................... | .................... |
| U | 1775 | hCoV-19/England/204781664/2020\|EPI_ISL_683568\|2020 | .................... | .................... | .................... | .................... | .................... | .................... |
| U | 1776 | hCoV-19/England/204781665/2020\|EPI_ISL_683569\|2020 | .................... | .................... | .................... | .................... | .................... | .................... |
| U | 1777 | hCoV-19/England/204781667/2020\|EPI_ISL_683570\|2020 | .................... | .................... | .................... | .................... | .................... | .................... |
| U | 1778 | hCoV-19/England/204781669/2020\|EPI_ISL_683571\|2020 | .................... | .................... | .................... | .................... | .................... | .................... |
| U | 1779 | hCoV-19/England/204790156/2020\|EPI_ISL_683572\|2020 | .................... | .................... | .................... | .................... | .................... | .................... |
| U | 1780 | hCoV-19/England/204820068/2020\|EPI_ISL_683573\|2020 | .................... | .................... | .................... | .................... | .................... | .................... |
| U | 1781 | hCoV-19/England/204820099/2020\|EPI_ISL_683574\|2020 | .................... | .................... | .................... | .................... | .................... | .................... |
| U | 1782 | hCoV-19/England/204820101/2020\|EPI_ISL_683575\|2020 | .................... | .................... | .................... | .................... | .................... | .................... |
| U | 1783 | hCoV-19/England/204821548_1/2020\|EPI_ISL_683576\|20 | .................... | .................... | .................... | .................... | .................... | .................... |
| U | 1784 | hCoV-19/England/204840111/2020\|EPI_ISL_683577\|2020 | .................... | .................... | .................... | .................... | .................... | .................... |
| U | 1785 | hCoV-19/England/204841906/2020\|EPI_ISL_683578\|2020 | .................... | .................... | .................... | .................... | .................... | .................... |
| U | 1786 | hCoV-19/England/204841909/2020\|EPI_ISL_683579\|2020 | .................... | .................... | .................... | .................... | .................... | .................... |
| U | 1787 | hCoV-19/England/204841910/2020\|EPI_ISL_683580\|2020 | .................... | .................... | .................... | .................... | .................... | .................... |
| U | 1788 | hCoV-19/England/204841912/2020\|EPI_ISL_683581\|2020 | .................... | .................... | .................... | .................... | .................... | .................... |
| U | 1789 | hCoV-19/England/204841915/2020\|EPI_ISL_683582\|2020 | .................... | .................... | .................... | .................... | .................... | .................... |
| U | 1790 | hCoV-19/England/204841916/2020\|EPI_ISL_683583\|2020 | .................... | .................... | .................... | .................... | .................... | .................... |
| U | 1791 | hCoV-19/England/204841918/2020\|EPI_ISL_683584\|2020 | .................... | .................... | .................... | .................... | .................... | .................... |
| U | 1792 | hCoV-19/England/204841921/2020\|EPI_ISL_683585\|2020 | .................... | .................... | .................... | .................... | .................... | .................... |
| U | 1793 | hCoV-19/England/QEUH-BA33ED/2020\|EPI_ISL_672850\|20 | .................... | .................... | .................... | .................... | .................... | .................... |

| | | | | | | | |
|---|---|---|---|---|---|---|---|
| U | 1795 hCoV-19/England/QEUH-BA2E92/2020\|EPI_ISL_672853\|20 | ···················· | ···················· | ····················· | ···················· | ···················· | ···················· |
| U | 1796 hCoV-19/England/QEUH-BA2E1A/2020\|EPI_ISL_672854\|20 | ···················· | ···················· | ····················· | ···················· | ···················· | ···················· |
| U | 1799 hCoV-19/England/QEUH-BA1F18/2020\|EPI_ISL_672857\|20 | ···················· | ···················· | ····················· | ···················· | ···················· | ···················· |
| U | 1800 hCoV-19/England/QEUH-BA3189/2020\|EPI_ISL_672858\|20 | ···················· | ···················· | ····················· | ···················· | ···················· | ···················· |
| U | 1801 hCoV-19/England/QEUH-BA29A0/2020\|EPI_ISL_672859\|20 | ···················· | ···················· | ····················· | ···················· | ···················· | ···················· |
| U | 1802 hCoV-19/England/QEUH-BA116D/2020\|EPI_ISL_672860\|20 | ···················· | ···················· | ····················· | ···················· | ···················· | ···················· |
| U | 1803 hCoV-19/England/QEUH-BA1E84/2020\|EPI_ISL_672861\|20 | ···················· | ···················· | ····················· | ···················· | ···················· | ···················· |
| U | 1804 hCoV-19/England/QEUH-BA30F5/2020\|EPI_ISL_672863\|20 | ···················· | ···················· | ····················· | ···················· | ···················· | ···················· |
| U | 1805 hCoV-19/England/QEUH-BA2F9F/2020\|EPI_ISL_672864\|20 | ···················· | ···················· | ····················· | ···················· | ···················· | ···················· |
| U | 1806 hCoV-19/England/QEUH-BA1297/2020\|EPI_ISL_672866\|20 | ···················· | ···················· | ····················· | ···················· | ···················· | ···················· |
| U | 1807 hCoV-19/England/QEUH-BA2FAE/2020\|EPI_ISL_672867\|20 | ···················· | ···················· | ····················· | ···················· | ···················· | ···················· |
| U | 1808 hCoV-19/England/QEUH-BA2F53/2020\|EPI_ISL_672868\|20 | ···················· | ···················· | ····················· | ···················· | ···················· | ···················· |
| U | 1809 hCoV-19/England/QEUH-BA11B8/2020\|EPI_ISL_672869\|20 | ···················· | ···················· | ····················· | ···················· | ···················· | ···················· |
| U | 1810 hCoV-19/England/QEUH-BA315C/2020\|EPI_ISL_672870\|20 | ···················· | ···················· | ····················· | ···················· | ···················· | ···················· |
| U | 1811 hCoV-19/England/QEUH-BA11C7/2020\|EPI_ISL_672872\|20 | ···················· | ···················· | ····················· | ···················· | ···················· | ···················· |
| U | 1812 hCoV-19/England/QEUH-BA2973/2020\|EPI_ISL_672874\|20 | ···················· | ···················· | ····················· | ···················· | ···················· | ···················· |
| U | 1813 hCoV-19/England/QEUH-BA12F1/2020\|EPI_ISL_672876\|20 | ···················· | ···················· | ····················· | ···················· | ·····T·············· | ···················· |
| U | 1814 hCoV-19/England/QEUH-BA2EFC/2020\|EPI_ISL_672877\|20 | ···················· | ···················· | ····················· | ···················· | ···················· | ···················· |
| U | 1815 hCoV-19/England/QEUH-BA2F26/2020\|EPI_ISL_672878\|20 | ···················· | ···················· | ····················· | ···················· | ···················· | ···················· |
| U | 1816 hCoV-19/England/QEUH-BA3031/2020\|EPI_ISL_672879\|20 | ···················· | ···················· | ····················· | ···················· | ···················· | ···················· |
| U | 1817 hCoV-19/England/QEUH-BA1E48/2020\|EPI_ISL_672881\|20 | ···················· | ···················· | ····················· | ···················· | ···················· | ···················· |
| U | 1819 hCoV-19/England/QEUH-BA1EA2/2020\|EPI_ISL_672884\|20 | ···················· | ···················· | ····················· | ···················· | ···················· | ···················· |
| U | 1820 hCoV-19/England/QEUH-BA1EEE/2020\|EPI_ISL_672886\|20 | ···················· | ···················· | ····················· | ···················· | ···················· | ···················· |
| U | 1822 hCoV-19/England/QEUH-BA3417/2020\|EPI_ISL_672889\|20 | ···················· | ···················· | ····················· | ···················· | ·····T·············· | ···················· |
| U | 1823 hCoV-19/England/QEUH-BA1349/2020\|EPI_ISL_672893\|20 | ···················· | ···················· | ····················· | ···················· | ···················· | ···················· |
| U | 1824 hCoV-19/England/QEUH-BA2B8C/2020\|EPI_ISL_672894\|20 | ···················· | ···················· | ····················· | ···················· | ···················· | ···················· |
| U | 1825 hCoV-19/England/QEUH-BA29CE/2020\|EPI_ISL_672895\|20 | ···················· | ···················· | ····················· | ···················· | ···················· | ···················· |
| U | 1826 hCoV-19/England/QEUH-BA31C5/2020\|EPI_ISL_672897\|20 | ···················· | ···················· | ····················· | ···················· | ···················· | ···················· |
| U | 1827 hCoV-19/England/QEUH-BA10F7/2020\|EPI_ISL_672899\|20 | ···················· | ···················· | ····················· | ···················· | ···················· | ···················· |
| U | 1828 hCoV-19/England/CAMC-B97F9D/2020\|EPI_ISL_675039\|20 | ···················· | ···················· | ····················· | ···················· | ···················· | ···················· |
| U | 1829 hCoV-19/England/CAMC-B981E1/2020\|EPI_ISL_675040\|20 | ···················· | ···················· | ····················· | ···················· | ···················· | ···················· |
| U | 1830 hCoV-19/England/MILK-B977FF/2020\|EPI_ISL_675041\|20 | ···················· | ···················· | ····················· | ···················· | ···················· | ···················· |
| U | 1831 hCoV-19/England/CAMC-B982FD/2020\|EPI_ISL_675042\|20 | ···················· | ···················· | ····················· | ···················· | ·····T·············· | ···················· |
| U | 1832 hCoV-19/England/MILK-B9623B/2020\|EPI_ISL_675043\|20 | ···················· | ···················· | ····················· | ···················· | ···················· | ···················· |
| U | 1833 hCoV-19/England/MILK-B971A6/2020\|EPI_ISL_675045\|20 | ···················· | ···················· | ····················· | ···················· | ···················· | ···················· |
| U | 1834 hCoV-19/England/CAMC-B98345/2020\|EPI_ISL_675046\|20 | ···················· | ·A·················· | ····················· | ···················· | ···················· | ···················· |
| U | 1835 hCoV-19/England/MILK-B96F43/2020\|EPI_ISL_675047\|20 | ···················· | ···················· | ····················· | ···················· | ···················· | ···················· |
| U | 1836 hCoV-19/England/MILK-B96FE9/2020\|EPI_ISL_675048\|20 | ···················· | ···················· | ····················· | ···················· | ···················· | ···················· |
| U | 1837 hCoV-19/England/MILK-B95CF2/2020\|EPI_ISL_675049\|20 | ···················· | ···················· | ····················· | ···················· | ···················· | ···················· |
| U | 1838 hCoV-19/Scotland/QEUH-B6C887/2020\|EPI_ISL_676360\|2 | ···················· | ···················· | ····················· | ···················· | ···················· | ···················· |
| U | 1839 hCoV-19/Scotland/QEUH-B69723/2020\|EPI_ISL_676362\|2 | ···················· | ···················· | ····················· | ···················· | ···················· | ···················· |
| U | 1840 hCoV-19/Scotland/QEUH-B68BCE/2020\|EPI_ISL_676370\|2 | ···················· | ···················· | ····················· | ···················· | ···················· | ···················· |
| U | 1841 hCoV-19/Scotland/QEUH-B696F9/2020\|EPI_ISL_676371\|2 | ···················· | ···················· | ····················· | ···················· | ···················· | ···················· |
| U | 1842 hCoV-19/Scotland/QEUH-B68117/2020\|EPI_ISL_676372\|2 | ···················· | ···················· | ····················· | ···················· | ···················· | ···················· |
| U | 1843 hCoV-19/Scotland/QEUH-B65CDD/2020\|EPI_ISL_676374\|2 | ···················· | ···················· | ····················· | ···················· | ···················· | ···················· |
| U | 1844 hCoV-19/Scotland/QEUH-B696AE/2020\|EPI_ISL_676375\|2 | ···················· | ···················· | ····················· | ···················· | ···················· | ···················· |
| U | 1845 hCoV-19/Scotland/QEUH-B68C25/2020\|EPI_ISL_676376\|2 | ···················· | ···················· | ····················· | ···················· | ···················· | ···················· |
| U | 1846 hCoV-19/Scotland/QEUH-B696DB/2020\|EPI_ISL_676377\|2 | ···················· | ···················· | ····················· | ···················· | ···················· | ···················· |
| U | 1847 hCoV-19/Scotland/QEUH-B680A1/2020\|EPI_ISL_676378\|2 | ···················· | ···················· | ····················· | ···················· | ···················· | ···················· |
| U | 1848 hCoV-19/Scotland/QEUH-B69662/2020\|EPI_ISL_676379\|2 | ···················· | ···················· | ····················· | ···················· | ···················· | ···················· |
| U | 1849 hCoV-19/Scotland/QEUH-B2CE36/2020\|EPI_ISL_676400\|2 | ···················· | ···················· | ····················· | ···················· | ···················· | ···················· |
| U | 1850 hCoV-19/Scotland/QEUH-B2CC69/2020\|EPI_ISL_676401\|2 | ···················· | ···················· | ····················· | ···················· | ···················· | ···················· |
| U | 1851 hCoV-19/Scotland/QEUH-B2EE52/2020\|EPI_ISL_676402\|2 | ···················· | ···················· | ····················· | ···················· | ·····T·············· | ···················· |
| U | 1852 hCoV-19/Scotland/QEUH-B2CD0C/2020\|EPI_ISL_676403\|2 | ···················· | ···················· | ····················· | ···················· | ···················· | ···················· |
| U | 1853 hCoV-19/Scotland/QEUH-B2CD66/2020\|EPI_ISL_676404\|2 | ···················· | ···················· | ····················· | ···················· | ···················· | ···················· |
| U | 1854 hCoV-19/Scotland/QEUH-B2D2FD/2020\|EPI_ISL_676405\|2 | ···················· | ···················· | ····················· | ···················· | ···················· | ···················· |
| U | 1855 hCoV-19/Scotland/QEUH-B31830/2020\|EPI_ISL_676406\|2 | ···················· | ···················· | ····················· | ···················· | ···················· | ···················· |
| U | 1856 hCoV-19/Scotland/QEUH-B31812/2020\|EPI_ISL_676409\|2 | ···················· | ···················· | ····················· | ···················· | ···················· | ···················· |
| U | 1857 hCoV-19/Scotland/QEUH-B317D9/2020\|EPI_ISL_676410\|2 | ···················· | ···················· | ····················· | ···················· | ···················· | ···················· |
| U | 1858 hCoV-19/Scotland/QEUH-B31751/2020\|EPI_ISL_676411\|2 | ···················· | ···················· | ····················· | ···················· | ···················· | ···················· |
| U | 1859 hCoV-19/Scotland/QEUH-B317AC/2020\|EPI_ISL_676412\|2 | ···················· | ···················· | ····················· | ···················· | ···················· | ···················· |
| U | 1860 hCoV-19/Scotland/QEUH-B310FC/2020\|EPI_ISL_676413\|2 | ···················· | ···················· | ····················· | ···················· | ···················· | ···················· |
| U | 1861 hCoV-19/North Macedonia/2402/2020\|EPI_ISL_677700\|2 | ···················· | ···················· | ····················· | ···················· | ···················· | ···················· |
| U | 1862 hCoV-19/North Macedonia/2604/2020\|EPI_ISL_677701\|2 | ···················· | ···················· | ····················· | ···················· | ···················· | ···················· |
| U | 1863 hCoV-19/North Macedonia/3006/2020\|EPI_ISL_677702\|2 | ···················· | ···················· | ····················· | ···················· | ···················· | ···················· |
| U | 1864 hCoV-19/North Macedonia/3021/2020\|EPI_ISL_677703\|2 | ···················· | ···················· | ····················· | ···················· | ···················· | ···················· |
| U | 1865 hCoV-19/North Macedonia/3032/2020\|EPI_ISL_677704\|2 | ···················· | ···················· | ····················· | ···················· | ···················· | ···················· |
| U | 1866 hCoV-19/North Macedonia/3053/2020\|EPI_ISL_677705\|2 | ···················· | ···················· | ····················· | ···················· | ···················· | ···················· |
| U | 1867 hCoV-19/North Macedonia/3101/2020\|EPI_ISL_677706\|2 | ···················· | ···················· | ····················· | ···················· | ···················· | ···················· |
| U | 1868 hCoV-19/North Macedonia/3515/2020\|EPI_ISL_677707\|2 | ···················· | ···················· | ····················· | ···················· | ···················· | ···················· |
| U | 1869 hCoV-19/North Macedonia/4098/2020\|EPI_ISL_677708\|2 | ···················· | ···G················ | ····················· | ···················· | ···················· | ···················· |
| U | 1870 hCoV-19/North Macedonia/4637/2020\|EPI_ISL_677709\|2 | ···················· | ···················· | ····················· | ···················· | ···················· | ···················· |
| U | 1871 hCoV-19/North Macedonia/4668/2020\|EPI_ISL_677710\|2 | ···················· | ···················· | ····················· | ···················· | ···················· | ···················· |
| U | 1872 hCoV-19/North Macedonia/5003/2020\|EPI_ISL_677711\|2 | ···················· | ···················· | ····················· | ···················· | ···················· | ···················· |
| U | 1873 hCoV-19/North Macedonia/5047/2020\|EPI_ISL_677712\|2 | ···················· | ···················· | ····················· | ···················· | ···················· | ···················· |
| U | 1874 hCoV-19/North Macedonia/5312/2020\|EPI_ISL_677713\|2 | ···················· | ···················· | ····················· | ···················· | ···················· | ···················· |
| U | 1875 hCoV-19/North Macedonia/5313/2020\|EPI_ISL_677714\|2 | ···················· | ···················· | ····················· | ···················· | ···················· | ···················· |
| U | 1876 hCoV-19/North Macedonia/6752/2020\|EPI_ISL_677715\|2 | ···················· | ···················· | ····················· | ···················· | ···················· | ···················· |
| U | 1877 hCoV-19/North Macedonia/7238/2020\|EPI_ISL_677716\|2 | ···················· | ···················· | ····················· | ···················· | ···················· | ···················· |
| U | 1878 hCoV-19/North Macedonia/7813/2020\|EPI_ISL_677717\|2 | ···················· | ···················· | ····················· | ···················· | ···················· | ···················· |

```
U    1879 hCoV-19/North Macedonia/8000/2020|EPI_ISL_677718|2    ····················  ····················  ·····················  ····················  ····················  ····················
U    1880 hCoV-19/North Macedonia/8817/2020|EPI_ISL_677719|2    ····················  ····················  ·····················  ····················  ····················  ····················
U    1881 hCoV-19/North Macedonia/8867/2020|EPI_ISL_677720|2    ····················  ····················  ·····················  ····················  ····················  ····················
U    1882 hCoV-19/North Macedonia/8881/2020|EPI_ISL_677721|2    ····················  ····················  ·····················  ····················  ····················  ····················
U    1883 hCoV-19/North Macedonia/9073/2020|EPI_ISL_677722|2    ····················  ····················  ·····················  ····················  ····················  ····················
U    1884 hCoV-19/North Macedonia/9133/2020|EPI_ISL_677723|2    ····················  ····················  ·····················  ····················  ····················  ····················
U    1885 hCoV-19/North Macedonia/9304/2020|EPI_ISL_677724|2    ····················  ····················  ·····················  ····················  ····················  ····················
U    1886 hCoV-19/North Macedonia/9359/2020|EPI_ISL_677725|2    ····················  ····················  ·····················  ····················  ····················  ····················
U    1887 hCoV-19/North Macedonia/9360/2020|EPI_ISL_677726|2    ····················  ····················  ·····················  ····················  ····················  ····················
U    1888 hCoV-19/Hungary/US-60441w/2020|EPI_ISL_677727|2020    ····················  ····················  ·····················  ····················  ····················  ····················
U    1889 hCoV-19/Hungary/US-30275w/2020|EPI_ISL_677729|2020    ····················  ····················  ·····················  ····················  ····················  ····················
U    1890 hCoV-19/Hungary/US-30398w/2020|EPI_ISL_677730|2020    ····················  ····················  ·····················  ····················  ····················  ····················
U    1891 hCoV-19/Hungary/US-31847w/2020|EPI_ISL_677735|2020    ····················  ····················  ·····················  ····················  ····················  ····················
U    1892 hCoV-19/Hungary/US-31848w/2020|EPI_ISL_677736|2020    ····················  ····················  ·····················  ····················  ····················  ····················
U    1893 hCoV-19/Hungary/US-31849w/2020|EPI_ISL_677737|2020    ····················  ····················  ·····················  ····················  ····················  ····················
U    1894 hCoV-19/Hungary/US-32269w/2020|EPI_ISL_677739|2020    ····················  ····················  ·····················  ····················  ····················  ····················
U    1895 hCoV-19/Hungary/US-32294w/2020|EPI_ISL_677741|2020    ····················  ····················  ·····················  ····················  ····················  ····················
U    1896 hCoV-19/Hungary/US-32297w/2020|EPI_ISL_677742|2020    ····················  ····················  ·····················  ····················  ····················  ····················
U    1897 hCoV-19/Hungary/US-32312w/2020|EPI_ISL_677744|2020    ····················  ····················  ·····················  ····················  ····················  ····················
U    1898 hCoV-19/USA/MI-UM-10036767162/2020|EPI_ISL_682200|    ········T···········  ·············A······  ···········T·········  ····················  ····················  ····················
U    1902 hCoV-19/USA/MI-UM-10036747334/2020|EPI_ISL_682204|    ····················  ·············A······  ·····················  ····················  ····················  ····················
U    1903 hCoV-19/USA/MI-UM-10036767440/2020|EPI_ISL_682205|    ····················  ····················  ·····················  ····················  ····················  ····················
U    1904 hCoV-19/USA/MI-UM-10036742479/2020|EPI_ISL_682206|    ····················  ····················  ·····················  ····················  ····················  ····················
U    1905 hCoV-19/USA/MI-UM-10036767463/2020|EPI_ISL_682207|    ····················  ····················  ·····················  ····················  ····················  ····················
U    1906 hCoV-19/USA/MI-UM-10036746097/2020|EPI_ISL_682208|    ····················  ····················  ·····················  ····················  ····················  ····················
U    1914 hCoV-19/USA/MI-UM-10036755381/2020|EPI_ISL_682217|    ····················  ····················  ·····················  ····················  ····················  ····················
U    1916 hCoV-19/USA/MI-UM-10036767331/2020|EPI_ISL_682220|    ····················  ····················  ·····················  ····················  ····················  ····················
U    1917 hCoV-19/USA/MI-UM-10036762197/2020|EPI_ISL_682221|    ····················  ····················  ·····················  ····················  ····················  ····················
U    1918 hCoV-19/USA/MI-UM-10036759541/2020|EPI_ISL_682222|    ····················  ·A··················  ·····················  ····················  ····················  ····················
U    1920 hCoV-19/USA/MI-UM-10036765823/2020|EPI_ISL_682224|    ····················  ····················  ·····················  ····················  ····················  ····················
U    1921 hCoV-19/USA/MI-UM-10036767342/2020|EPI_ISL_682225|    ····················  ····················  ·····················  ····················  ····················  ····················
U    1923 hCoV-19/USA/MI-UM-10036758028/2020|EPI_ISL_682227|    ····················  ····················  ·····················  ····················  ····················  ····················
U    1924 hCoV-19/USA/MI-UM-10036765841/2020|EPI_ISL_682228|    ····················  ····················  ·····················  ····················  ····················  ····················
U    1925 hCoV-19/USA/MI-UM-10036767316/2020|EPI_ISL_682229|    ····················  ·············A······  ·····················  ····················  ····················  ····················
U    1929 hCoV-19/Costa Rica/INC-0093/2020|EPI_ISL_682235|20    ····················  ····················  ·····················  ····················  ····················  ····················
U    1930 hCoV-19/Costa Rica/INC-0094/2020|EPI_ISL_682236|20    ····················  ····················  ·····················  ····················  ····················  ····················
U    1931 hCoV-19/Costa Rica/INC-0096/2020|EPI_ISL_682238|20    ····················  ····················  ·····················  ····················  ····················  ····················
U    1932 hCoV-19/Costa Rica/INC-0097/2020|EPI_ISL_682239|20    ····················  ····················  ·····················  ····················  ····················  ····················
U    1933 hCoV-19/Costa Rica/INC-0115/2020|EPI_ISL_682255|20    ····················  ····················  ·····················  ····················  ····················  ····················
U    1934 hCoV-19/Costa Rica/INC-0116/2020|EPI_ISL_682256|20    ····················  ····················  ·····················  ····················  ····················  ····················
U    1935 hCoV-19/Costa Rica/INC-0118/2020|EPI_ISL_682257|20    ····················  ····················  ·····················  ····················  ····················  ····················
U    1936 hCoV-19/Costa Rica/INC-0119/2020|EPI_ISL_682258|20    ····················  ····················  ·····················  ····················  ····················  ····················
U    1937 hCoV-19/Costa Rica/INC-0120/2020|EPI_ISL_682259|20    ····················  ····················  ·····················  ····················  ····················  ····················
U    1938 hCoV-19/Costa Rica/INC-0121/2020|EPI_ISL_682260|20    ····················  ····················  ·····················  ····················  ····················  ····················
U    1939 hCoV-19/Costa Rica/INC-0122/2020|EPI_ISL_682261|20    ····················  ····················  ·····················  ····················  ····················  ····················
U    1940 hCoV-19/Costa Rica/INC-0123/2020|EPI_ISL_682262|20    ····················  ····················  ·····················  ····················  ····················  ····················
U    1941 hCoV-19/Costa Rica/INC-0124/2020|EPI_ISL_682263|20    ····················  ····················  ·····················  ····················  ····················  ····················
U    1942 hCoV-19/Costa Rica/INC-0125/2020|EPI_ISL_682264|20    ····················  ····················  ·····················  ····················  ····················  ····················
U    1943 hCoV-19/Costa Rica/INC-0126/2020|EPI_ISL_682265|20    ····················  ····················  ·····················  ····················  ····················  ····················
U    1944 hCoV-19/Costa Rica/INC-0127/2020|EPI_ISL_682266|20    ····················  ····················  ·····················  ····················  ····················  ····················
U    1945 hCoV-19/Costa Rica/INC-0128/2020|EPI_ISL_682267|20    ····················  ····················  ·····················  ····················  ····················  ····················
U    1946 hCoV-19/Costa Rica/INC-0129/2020|EPI_ISL_682268|20    ····················  ····················  ·····················  ····················  ····················  ····················
U    1947 hCoV-19/Costa Rica/INC-0130/2020|EPI_ISL_682269|20    ····················  ····················  ·····················  ····················  ····················  ····················
U    1948 hCoV-19/Costa Rica/INC-0131/2020|EPI_ISL_682270|20    ····················  ····················  ·····················  ····················  ····················  ····················
U    1949 hCoV-19/Costa Rica/INC-0132/2020|EPI_ISL_682271|20    ····················  ····················  ·····················  ····················  ····················  ····················
U    1950 hCoV-19/Costa Rica/INC-0133/2020|EPI_ISL_682272|20    ····················  ····················  ·····················  ····················  ····················  ····················
U    1951 hCoV-19/Costa Rica/INC-0135/2020|EPI_ISL_682273|20    ····················  ····················  ·····················  ····················  ····················  ····················
U    1952 hCoV-19/Costa Rica/INC-0137/2020|EPI_ISL_682275|20    ····················  ····················  ·····················  ····················  ····················  ····················
U    1953 hCoV-19/Costa Rica/INC-0138/2020|EPI_ISL_682276|20    ····················  ····················  ·····················  ····················  ····················  ····················
U    1954 hCoV-19/Costa Rica/INC-0139/2020|EPI_ISL_682277|20    ····················  ····················  ·····················  ····················  ····················  ····················
U    1955 hCoV-19/New Zealand/20CV0598/2020|EPI_ISL_682279|2    ····················  ·A··················  ·····················  ····················  ····················  ····················
U    1956 hCoV-19/New Zealand/20CV0599/2020|EPI_ISL_682280|2    ····················  ····················  ·····················  ····················  ····················  ····················
U    1957 hCoV-19/New Zealand/20CV0600/2020|EPI_ISL_682281|2    ····················  ····················  ·····················  ····················  ····················  ····················
U    1958 hCoV-19/New Zealand/20CV0601/2020|EPI_ISL_682282|2    ····················  ····················  ·····················  ····················  ····················  ····················
U    1959 hCoV-19/New Zealand/20CV0604/2020|EPI_ISL_682283|2    ····················  ····················  ·····················  ····················  ····················  ····················
U    1960 hCoV-19/New Zealand/20CV0607/2020|EPI_ISL_682286|2    ····················  ····················  ·····················  ····················  ····················  ····················
U    1961 hCoV-19/New Zealand/20CV0608/2020|EPI_ISL_682287|2    ····················  ····················  ·····················  ····················  ····················  ····················
U    1962 hCoV-19/New Zealand/20CV0610/2020|EPI_ISL_682288|2    ····················  ····················  ·····················  ····················  ····················  ····················
U    1963 hCoV-19/New Zealand/20CV0611/2020|EPI_ISL_682289|2    ····················  ····················  ·····················  ····················  ····················  ····················
U    1964 hCoV-19/New Zealand/20CV0612/2020|EPI_ISL_682290|2    ····················  ····················  ·····················  ····················  ····················  ····················
U    1965 hCoV-19/New Zealand/20CV0613/2020|EPI_ISL_682291|2    ····················  ····················  ·····················  ····················  ····················  ····················
U    1966 hCoV-19/New Zealand/20CV0614/2020|EPI_ISL_682292|2    ····················  ····················  ·····················  ····················  ····················  ····················
U    1967 hCoV-19/New Zealand/20CV0615/2020|EPI_ISL_682293|2    ····················  ····················  ·····················  ····················  ····················  ····················
U    1968 hCoV-19/New Zealand/20CV0617/2020|EPI_ISL_682294|2    ····················  ····················  ·····················  ····················  ····················  ····················
U    1969 hCoV-19/New Zealand/20CV0619/2020|EPI_ISL_682295|2    ····················  ····················  ·····················  ····················  ····················  ····················
U    1970 hCoV-19/New Zealand/20CV0625/2020|EPI_ISL_682296|2    ····················  ····················  ·····················  ····················  ····················  ····················
U    1971 hCoV-19/Cambodia/Kunming_kms-2/2020|EPI_ISL_682298    ····················  ····················  ·····················  ····················  ····················  ····················
U    1972 hCoV-19/Bahrain/340957986/2020|EPI_ISL_682299|2020    ····················  ····················  ·····················  ····················  ····················  ····················
U    1973 hCoV-19/England/204590456/2020|EPI_ISL_683500|2020    ····················  ····················  ·····················  ····················  ····················  ····················
U    1974 hCoV-19/England/204590457/2020|EPI_ISL_683501|2020    ····················  ····················  ·····················  ····················  ····················  ····················
```

| | | | | | | | |
|---|---|---|---|---|---|---|---|
| U | 1975 hCoV-19/England/204590460/2020\|EPI_ISL_683502\|2020 | ···················· | ···················· | ···················· | ···················· | ···················· | ···················· |
| U | 1976 hCoV-19/England/204590461/2020\|EPI_ISL_683503\|2020 | ···················· | ···················· | ···················· | ···················· | ···················· | ···················· |
| U | 1977 hCoV-19/England/204590463/2020\|EPI_ISL_683504\|2020 | ···················· | ···················· | ···················· | ···················· | ···················· | ···················· |
| U | 1978 hCoV-19/England/204590464/2020\|EPI_ISL_683505\|2020 | ···················· | ···················· | ···················· | ···················· | ···················· | ···················· |
| U | 1979 hCoV-19/England/204590465/2020\|EPI_ISL_683506\|2020 | ···················· | ···················· | ···················· | ···················· | ···················· | ···················· |
| U | 1980 hCoV-19/England/204590466/2020\|EPI_ISL_683507\|2020 | ···················· | ···················· | ···················· | ···················· | ···················· | ···················· |
| U | 1981 hCoV-19/England/204590480/2020\|EPI_ISL_683508\|2020 | ···················· | ···················· | ···················· | ···················· | ···················· | ···················· |
| U | 1982 hCoV-19/England/204590483/2020\|EPI_ISL_683509\|2020 | ···················· | ···················· | ···················· | ···················· | ···················· | ···················· |
| U | 1983 hCoV-19/England/204590485/2020\|EPI_ISL_683510\|2020 | ···················· | ···················· | ···················· | ···················· | ···················· | ···················· |
| U | 1984 hCoV-19/England/204590488/2020\|EPI_ISL_683511\|2020 | ···················· | ···················· | ···················· | ···················· | ···················· | ···················· |
| U | 1985 hCoV-19/England/204590491/2020\|EPI_ISL_683512\|2020 | ···················· | ···················· | ···················· | ···················· | ···················· | ···················· |
| U | 1986 hCoV-19/England/204590493/2020\|EPI_ISL_683513\|2020 | ···················· | ···················· | ···················· | ···················· | ···················· | ···················· |
| U | 1987 hCoV-19/England/204590500/2020\|EPI_ISL_683514\|2020 | ···················· | ···················· | ···················· | ···················· | ···················· | ···················· |
| U | 1988 hCoV-19/England/204590503/2020\|EPI_ISL_683515\|2020 | ···················· | ···················· | ···················· | ···················· | ···················· | ···················· |
| U | 1989 hCoV-19/England/204642472/2020\|EPI_ISL_683516\|2020 | ···················· | ···················· | ···················· | ···················· | ······T············· | ···················· |
| U | 1990 hCoV-19/England/204642473/2020\|EPI_ISL_683517\|2020 | ···················· | ···················· | ···················· | ···················· | ······T············· | ···················· |
| U | 1991 hCoV-19/England/204661404/2020\|EPI_ISL_683518\|2020 | ···················· | ···················· | ···················· | ···················· | ······T············· | ···················· |
| U | 1992 hCoV-19/England/204690073/2020\|EPI_ISL_683519\|2020 | ···················· | ···················· | ···················· | ···················· | ···················· | ···················· |
| U | 1993 hCoV-19/England/204690077/2020\|EPI_ISL_683520\|2020 | ···················· | ···················· | ···················· | ···················· | ···················· | ···················· |
| U | 1994 hCoV-19/England/204690078/2020\|EPI_ISL_683521\|2020 | ···················· | ···················· | ···················· | ···················· | ···················· | ···················· |
| U | 1995 hCoV-19/England/204690089/2020\|EPI_ISL_683522\|2020 | ···················· | ···················· | ···················· | ···················· | ···················· | ···················· |
| U | 1996 hCoV-19/England/204690091/2020\|EPI_ISL_683523\|2020 | ···················· | ···················· | ···················· | ···················· | ···················· | ···················· |
| U | 1997 hCoV-19/England/204690092/2020\|EPI_ISL_683524\|2020 | ···················· | ···················· | ···················· | ···················· | ···················· | ···················· |
| U | 1998 hCoV-19/England/204690093/2020\|EPI_ISL_683525\|2020 | ···················· | ···················· | ···················· | ···················· | ···················· | ···················· |
| U | 1999 hCoV-19/England/204690094/2020\|EPI_ISL_683526\|2020 | ···················· | ···················· | ···················· | ···················· | ···················· | ···················· |
| U | 2000 hCoV-19/England/204690096/2020\|EPI_ISL_683527\|2020 | ···················· | ···················· | ···················· | ···················· | ···················· | ···················· |
| U | 2001 hCoV-19/England/204690097/2020\|EPI_ISL_683528\|2020 | ···················· | ···················· | ···················· | ···················· | ···················· | ···················· |
| U | 2002 hCoV-19/England/204690098/2020\|EPI_ISL_683529\|2020 | ···················· | ···················· | ···················· | ···················· | ···················· | ···················· |
| U | 2003 hCoV-19/England/204690307/2020\|EPI_ISL_683530\|2020 | ···················· | ···················· | ···················· | ···················· | ···················· | ···················· |
| U | 2005 hCoV-19/England/204690328/2020\|EPI_ISL_683532\|2020 | ···················· | ···················· | ···················· | ···················· | ···················· | ···················· |
| U | 2006 hCoV-19/England/204690337/2020\|EPI_ISL_683533\|2020 | ···················· | ···················· | ···················· | ···················· | ···················· | ···················· |
| U | 2007 hCoV-19/England/204721133/2020\|EPI_ISL_683534\|2020 | ···················· | ···················· | ···················· | ···················· | ···················· | ···················· |
| U | 2008 hCoV-19/England/204721137/2020\|EPI_ISL_683535\|2020 | ···················· | ···················· | ···················· | ···················· | ···················· | ···················· |
| U | 2009 hCoV-19/England/204722542/2020\|EPI_ISL_683536\|2020 | ···················· | ···················· | ···················· | ···················· | ······T············· | ···················· |
| U | 2010 hCoV-19/England/204722544/2020\|EPI_ISL_683537\|2020 | ···················· | ···················· | ···················· | ···················· | ···················· | ···················· |
| U | 2011 hCoV-19/England/204742379/2020\|EPI_ISL_683538\|2020 | ···················· | ···················· | ···················· | ···················· | ···················· | ···················· |
| U | 2012 hCoV-19/England/204742475/2020\|EPI_ISL_683539\|2020 | ···················· | ···················· | ···················· | ···················· | ···················· | ···················· |
| U | 2013 hCoV-19/England/204742476/2020\|EPI_ISL_683540\|2020 | ···················· | ···················· | ···················· | ···················· | ···················· | ···················· |
| U | 2014 hCoV-19/England/204742477/2020\|EPI_ISL_683541\|2020 | ···················· | ···················· | ···················· | ···················· | ···················· | ···················· |
| U | 2015 hCoV-19/England/204742478/2020\|EPI_ISL_683542\|2020 | ···················· | ···················· | ···················· | ···················· | ···················· | ···················· |
| U | 2016 hCoV-19/England/204742480/2020\|EPI_ISL_683543\|2020 | ···················· | ···················· | ···················· | ···················· | ···················· | ···················· |
| U | 2017 hCoV-19/England/204742484/2020\|EPI_ISL_683544\|2020 | ···················· | ···················· | ···················· | ···················· | ···················· | ···················· |
| U | 2018 hCoV-19/England/204742485/2020\|EPI_ISL_683545\|2020 | ···················· | ···················· | ···················· | ···················· | ···················· | ···················· |
| U | 2019 hCoV-19/England/204760987/2020\|EPI_ISL_683546\|2020 | ···················· | ···················· | ···················· | ···················· | ···················· | ···················· |
| U | 2020 hCoV-19/England/204760990/2020\|EPI_ISL_683547\|2020 | ···················· | ···················· | ···················· | ···················· | ······T············· | ···················· |
| U | 2021 hCoV-19/England/204761663/2020\|EPI_ISL_683548\|2020 | ···················· | ···················· | ···················· | ···················· | ···················· | ···················· |
| U | 2022 hCoV-19/England/204761664/2020\|EPI_ISL_683549\|2020 | ···················· | ···················· | ···················· | ···················· | ···················· | ···················· |
| U | 2023 hCoV-19/England/204761665/2020\|EPI_ISL_683550\|2020 | ···················· | ···················· | ···················· | ···················· | ···················· | ···················· |
| U | 2024 hCoV-19/England/204761666/2020\|EPI_ISL_683551\|2020 | ···················· | ···················· | ···················· | ···················· | ···················· | ···················· |
| U | 2025 hCoV-19/England/204761667/2020\|EPI_ISL_683552\|2020 | ···················· | ···················· | ···················· | ···················· | ···················· | ···················· |
| U | 2026 hCoV-19/England/204761668/2020\|EPI_ISL_683553\|2020 | ···················· | ···················· | ···················· | ···················· | ···················· | ···················· |
| U | 2027 hCoV-19/England/204761673/2020\|EPI_ISL_683554\|2020 | ···················· | ···················· | ···················· | ···················· | ···················· | ···················· |
| U | 2028 hCoV-19/England/204761674/2020\|EPI_ISL_683555\|2020 | ···················· | ···················· | ···················· | ···················· | ···················· | ···················· |
| U | 2029 hCoV-19/England/204761675/2020\|EPI_ISL_683556\|2020 | ···················· | ···················· | ···················· | ···················· | ···················· | ···················· |
| U | 2030 hCoV-19/England/204761680/2020\|EPI_ISL_683557\|2020 | ···················· | ···················· | ···················· | ···················· | ···················· | ···················· |
| U | 2031 hCoV-19/England/204761681/2020\|EPI_ISL_683558\|2020 | ···················· | ···················· | ···················· | ···················· | ···················· | ···················· |
| U | 2032 hCoV-19/England/204761684/2020\|EPI_ISL_683559\|2020 | ···················· | ···················· | ···················· | ···················· | ···················· | ···················· |
| U | 2033 hCoV-19/England/204761685/2020\|EPI_ISL_683560\|2020 | ···················· | ···················· | ···················· | ···················· | ···················· | ···················· |
| U | 2034 hCoV-19/England/204761686/2020\|EPI_ISL_683561\|2020 | ···················· | ···················· | ···················· | ···················· | ···················· | ···················· |
| U | 2035 hCoV-19/England/204762042/2020\|EPI_ISL_683562\|2020 | ···················· | ···················· | ···················· | ···················· | ···················· | ···················· |
| U | 2036 hCoV-19/England/204780331/2020\|EPI_ISL_683563\|2020 | ···················· | ···················· | ···················· | ···················· | ···················· | ···················· |
| U | 2037 hCoV-19/England/204780433/2020\|EPI_ISL_683564\|2020 | ···················· | ···················· | ···················· | ···················· | ···················· | ···················· |
| U | 2038 hCoV-19/England/204780434/2020\|EPI_ISL_683565\|2020 | ···················· | ···················· | ···················· | ···················· | ···················· | ···················· |
| U | 2039 hCoV-19/England/204781278/2020\|EPI_ISL_683566\|2020 | ···················· | ···················· | ···················· | ···················· | ······T············· | ···················· |
| U | 2040 hCoV-19/England/204781283/2020\|EPI_ISL_683567\|2020 | ···················· | ···················· | ···················· | ···················· | ······T············· | ···················· |
| U | 2041 hCoV-19/England/204841926/2020\|EPI_ISL_683586\|2020 | ···················· | ···················· | ···················· | ···················· | ···················· | ···················· |
| U | 2042 hCoV-19/England/204841928/2020\|EPI_ISL_683587\|2020 | ···················· | ···················· | ···················· | ···················· | ···················· | ···················· |
| U | 2043 hCoV-19/England/204841929/2020\|EPI_ISL_683588\|2020 | ···················· | ···················· | ···················· | ···················· | ···················· | ···················· |
| U | 2044 hCoV-19/England/204841931/2020\|EPI_ISL_683589\|2020 | ···················· | ···················· | ···················· | ···················· | ···················· | ···················· |
| U | 2045 hCoV-19/England/204860245/2020\|EPI_ISL_683590\|2020 | ···················· | ···················· | ···················· | ···················· | ······T············· | ···················· |
| U | 2046 hCoV-19/England/204860406/2020\|EPI_ISL_683591\|2020 | ···················· | ···················· | ···················· | ···················· | ···················· | ···················· |
| U | 2050 hCoV-19/England/QEUH-BA2B7D/2020\|EPI_ISL_672796\|20 | ···················· | ···················· | ···················· | ···················· | ···················· | ···················· |
| U | 2051 hCoV-19/England/QEUH-BA29EC/2020\|EPI_ISL_672797\|20 | ···················· | ···················· | ···················· | ···················· | ···················· | ···················· |
| U | 2052 hCoV-19/England/QEUH-BA268B/2020\|EPI_ISL_672798\|20 | ···················· | ···················· | ···················· | ···················· | ···················· | ···················· |
| U | 2053 hCoV-19/England/QEUH-BA1F36/2020\|EPI_ISL_672799\|20 | ···················· | ···················· | ···················· | ···················· | ···················· | ···················· |
| U | 2054 hCoV-19/England/QEUH-BA2630/2020\|EPI_ISL_672804\|20 | ···················· | ···················· | ···················· | ···················· | ···················· | ···················· |
| U | 2055 hCoV-19/England/QEUH-BA13A3/2020\|EPI_ISL_672805\|20 | ···················· | ···················· | ···················· | ···················· | ···················· | ···················· |
| U | 2056 hCoV-19/England/QEUH-BA3444/2020\|EPI_ISL_672807\|20 | ···················· | ···················· | ···················· | ···················· | ···················· | ···················· |
| U | 2057 hCoV-19/England/QEUH-BA121F/2020\|EPI_ISL_672810\|20 | ···················· | ···················· | ···················· | ···················· | ···················· | ···················· |
| U | 2058 hCoV-19/England/QEUH-BA2F80/2020\|EPI_ISL_672811\|20 | ···················· | ···················· | ···················· | ···················· | ···················· | ···················· |

U 2059 hCoV-19/England/QEUH-BA2A25/2020|EPI_ISL_672813|20 ···················· ···················· ····················· ···················· ···················· ····················
U 2060 hCoV-19/England/QEUH-BA10D9/2020|EPI_ISL_672819|20 ···················· ···················· ····················· ···················· ···················· ····················
U 2061 hCoV-19/England/QEUH-BA1EB1/2020|EPI_ISL_672823|20 ···················· ···················· ····················· ···················· ···················· ····················
U 2062 hCoV-19/England/QEUH-BA29DD/2020|EPI_ISL_672824|20 ···················· ···················· ····················· ···················· ···················· ····················
U 2063 hCoV-19/England/QEUH-BA115E/2020|EPI_ISL_672825|20 ···················· ···················· ····················· ···················· ···················· ····················
U 2064 hCoV-19/England/QEUH-BA125B/2020|EPI_ISL_672828|20 ···················· ···················· ····················· ···················· ···················· ····················
U 2065 hCoV-19/England/QEUH-BA266D/2020|EPI_ISL_672829|20 ···················· ···················· ····················· ···················· ···················· ····················
U 2066 hCoV-19/England/QEUH-BA2F62/2020|EPI_ISL_672834|20 ···················· ···················· ····················· ···················· ···················· ····················
U 2067 hCoV-19/England/QEUH-BA31A7/2020|EPI_ISL_672835|20 ···················· ···················· ····················· ···················· ···················· ····················
U 2068 hCoV-19/England/QEUH-BA317A/2020|EPI_ISL_672836|20 ···················· ···················· ····················· ···················· ···················· ····················
U 2069 hCoV-19/England/QEUH-BA3392/2020|EPI_ISL_672837|20 ···················· ···················· ····················· ···················· ···················· ····················
U 2070 hCoV-19/England/QEUH-BA124C/2020|EPI_ISL_672838|20 ···················· ···················· ····················· ···················· ···················· ····················
U 2071 hCoV-19/England/QEUH-BA29FB/2020|EPI_ISL_672840|20 ···················· ·A·················· ····················· ···················· ···················· ··A·················
U 2072 hCoV-19/England/QEUH-BA29BF/2020|EPI_ISL_672841|20 ···················· ···················· ····················· ···················· ···················· ····················
U 2073 hCoV-19/England/QEUH-BA0F46/2020|EPI_ISL_672844|20 ···················· ···················· ····················· ···················· ···················· ····················
U 2074 hCoV-19/England/QEUH-BA1EFD/2020|EPI_ISL_672845|20 ···················· ···················· ····················· ···················· ···················· ····················
U 2075 hCoV-19/England/QEUH-BA3408/2020|EPI_ISL_672846|20 ···················· ···················· ····················· ···················· ···················· ····················
U 2076 hCoV-19/England/QEUH-BA2EB0/2020|EPI_ISL_672848|20 ···················· ···················· ····················· ···················· ···················· ····················
U 2077 hCoV-19/England/MILK-B9620E/2020|EPI_ISL_675000|20 ···················· ···················· ····················· ···················· ···················· ····················
U 2078 hCoV-19/England/MILK-B95991/2020|EPI_ISL_675002|20 ···················· ···················· ····················· ···················· ···················· ····················
U 2079 hCoV-19/England/MILK-B961D4/2020|EPI_ISL_675003|20 ···················· ···················· ····················· ···················· ···················· ····················
U 2080 hCoV-19/England/MILK-B96CE2/2020|EPI_ISL_675005|20 ···················· ···················· ····················· ···················· ···················· ····················
U 2081 hCoV-19/England/MILK-B96DA3/2020|EPI_ISL_675007|20 ···················· ···················· ····················· ···················· ···················· ····················
U 2082 hCoV-19/England/CAMC-B980C6/2020|EPI_ISL_675008|20 ···················· ···················· ····················· ···················· ···················· ····················
U 2083 hCoV-19/England/MILK-B972B2/2020|EPI_ISL_675010|20 ···················· ···················· ····················· ···················· ···················· ····················
U 2084 hCoV-19/England/MILK-B97531/2020|EPI_ISL_675013|20 ···················· ···················· ····················· ···················· ···················· ····················
U 2085 hCoV-19/England/MILK-B970D6/2020|EPI_ISL_675014|20 ···················· ···················· ····················· ···················· ···················· ····················
U 2086 hCoV-19/England/MILK-B95CD4/2020|EPI_ISL_675015|20 ···················· ···················· ····················· ···················· ···················· ····················
U 2087 hCoV-19/England/MILK-B96D67/2020|EPI_ISL_675017|20 ···················· ···················· ····················· ···················· ···················· ····················
U 2088 hCoV-19/England/CAMC-B97F60/2020|EPI_ISL_675018|20 ···················· ···················· ····················· ···················· ···················· ····················
U 2089 hCoV-19/England/MILK-B95EA1/2020|EPI_ISL_675020|20 ···················· ···················· ····················· ···················· ···················· ····················
U 2090 hCoV-19/England/MILK-B977A4/2020|EPI_ISL_675022|20 ···················· ···················· ····················· ···················· ···················· ····················
U 2091 hCoV-19/England/MILK-B95B31/2020|EPI_ISL_675023|20 ···················· ·A·················· ····················· ···················· ···················· ··A·················
U 2092 hCoV-19/England/CAMC-B982C0/2020|EPI_ISL_675024|20 ···················· ···················· ····················· ···················· ···················· ····················
U 2093 hCoV-19/England/MILK-B97337/2020|EPI_ISL_675025|20 ···················· ···················· ····················· ···················· ···················· ····················
U 2094 hCoV-19/England/CAMC-B9866A/2020|EPI_ISL_675026|20 ···················· ···················· ····················· ···················· ···················· ····················
U 2095 hCoV-19/England/MILK-B9707C/2020|EPI_ISL_675028|20 ···················· ···················· ····················· ···················· ···················· ····················
U 2096 hCoV-19/England/MILK-B96295/2020|EPI_ISL_675029|20 ···················· ···················· ····················· ···················· ···················· ····················
U 2097 hCoV-19/England/MILK-B96F8F/2020|EPI_ISL_675030|20 ···················· ···················· ····················· ···················· ···················· ····················
U 2098 hCoV-19/England/MILK-B96E55/2020|EPI_ISL_675031|20 ···················· ···················· ····················· ···················· ···················· ····················
U 2099 hCoV-19/England/MILK-B97285/2020|EPI_ISL_675032|20 ···················· ···················· ····················· ···················· ···················· ····················
U 2100 hCoV-19/England/CAMC-B9813C/2020|EPI_ISL_675033|20 ···················· ·A·················· ····················· ···················· ···················· ··A·················
U 2101 hCoV-19/England/MILK-B96C4C/2020|EPI_ISL_675034|20 ···················· ···················· ····················· ···················· ···················· ····················
U 2102 hCoV-19/England/MILK-B96523/2020|EPI_ISL_675035|20 ···················· ···················· ····················· ···················· ···················· ····················
U 2103 hCoV-19/England/MILK-B96C97/2020|EPI_ISL_675036|20 ···················· ···················· ····················· ···················· ···················· ····················
U 2104 hCoV-19/England/MILK-B96480/2020|EPI_ISL_675037|20 ···················· ···················· ····················· ···················· ·····T·············· ····················
U 2105 hCoV-19/England/CAMC-B98248/2020|EPI_ISL_675038|20 ···················· ···················· ····················· ···················· ···················· ····················
U 2106 hCoV-19/England/MILK-B96F25/2020|EPI_ISL_675050|20 ···················· ·A·················· ····················· ···················· ···················· ··A·················
U 2107 hCoV-19/England/MILK-B95B40/2020|EPI_ISL_675051|20 ···················· ···················· ····················· ···················· ···················· ····················
U 2108 hCoV-19/England/MILK-B974E9/2020|EPI_ISL_675052|20 ···················· ···················· ····················· ···················· ···················· ····················
U 2109 hCoV-19/England/CAMC-B98275/2020|EPI_ISL_675053|20 ···················· ···················· ····················· ···················· ···················· ····················
U 2110 hCoV-19/England/MILK-B97504/2020|EPI_ISL_675054|20 ···················· ···················· ····················· ···················· ···················· ····················
U 2111 hCoV-19/England/MILK-B95CC5/2020|EPI_ISL_675055|20 ···················· ···················· ····················· ···················· ···················· ····················
U 2112 hCoV-19/England/MILK-B974F8/2020|EPI_ISL_675057|20 ···················· ···················· ····················· ···················· ···················· ····················
U 2113 hCoV-19/England/MILK-B961F2/2020|EPI_ISL_675060|20 ··················T· ···················· ····················· ···················· ·····T·············· ···················· 1
U 2114 hCoV-19/England/MILK-B96D49/2020|EPI_ISL_675061|20 ···················· ···················· ····················· ···················· ···················· ····················
U 2115 hCoV-19/England/MILK-B96BC7/2020|EPI_ISL_675062|20 ···················· ···················· ····················· ···················· ···················· ····················
U 2116 hCoV-19/England/MILK-B9766B/2020|EPI_ISL_675063|20 ···················· ···················· ····················· ···················· ···················· ····················
U 2117 hCoV-19/England/CAMC-B98020/2020|EPI_ISL_675064|20 ···················· ···················· ····················· ···················· ···················· ····················
U 2118 hCoV-19/England/MILK-B9722B/2020|EPI_ISL_675065|20 ···················· ···················· ····················· ···················· ···················· ····················
U 2119 hCoV-19/England/MILK-B96B5E/2020|EPI_ISL_675066|20 ···················· ···················· ····················· ···················· ···················· ····················
U 2120 hCoV-19/England/MILK-B95937/2020|EPI_ISL_675067|20 ···················· ···················· ····················· ···················· ···················· ····················
U 2121 hCoV-19/England/MILK-B95ECF/2020|EPI_ISL_675068|20 ···················· ···················· ····················· ···················· ···················· ····················
U 2122 hCoV-19/England/MILK-B9657E/2020|EPI_ISL_675069|20 ···················· ·········T·········· ····················· ···················· ···················· ····················
U 2123 hCoV-19/England/MILK-B975E6/2020|EPI_ISL_675070|20 ···················· ···················· ····················· ···················· ···················· ····················
U 2124 hCoV-19/England/CAMC-B9863D/2020|EPI_ISL_675071|20 ···················· ···················· ····················· ···················· ·····T·············· ····················
U 2125 hCoV-19/England/MILK-B95E65/2020|EPI_ISL_675072|20 ···················· ···················· ····················· ···················· ···················· ····················
U 2126 hCoV-19/England/MILK-B973BF/2020|EPI_ISL_675074|20 ···················· ···················· ····················· ···················· ···················· ····················
U 2127 hCoV-19/England/CAMC-B9803F/2020|EPI_ISL_675075|20 ···················· ···················· ····················· ···················· ···················· ····················
U 2128 hCoV-19/England/MILK-B96C88/2020|EPI_ISL_675076|20 ···················· ···················· ····················· ···················· ···················· ····················
U 2129 hCoV-19/England/MILK-B96CB5/2020|EPI_ISL_675077|20 ···················· ···················· ····················· ···················· ···················· ····················
U 2130 hCoV-19/England/MILK-B96C6A/2020|EPI_ISL_675078|20 ···················· ···················· ····················· ···················· ···················· ····················
U 2131 hCoV-19/England/MILK-B973CE/2020|EPI_ISL_675079|20 ···················· ···················· ····················· ···················· ···················· ····················
U 2132 hCoV-19/England/CAMC-B9821B/2020|EPI_ISL_675082|20 ···················· ···················· ····················· ···················· ···················· ····················
U 2133 hCoV-19/England/MILK-B9709A/2020|EPI_ISL_675083|20 ···················· ···················· ····················· ···················· ···················· ····················
U 2134 hCoV-19/England/MILK-B977B3/2020|EPI_ISL_675084|20 ···················· ···················· ····················· ···················· ·····T·············· ····················
U 2135 hCoV-19/England/MILK-B971C4/2020|EPI_ISL_675085|20 ···················· ···················· ····················· ···················· ···················· ····················
U 2136 hCoV-19/England/MILK-B9714C/2020|EPI_ISL_675086|20 ···················· ···················· ····················· ···················· ···················· ····················
U 2137 hCoV-19/England/CAMC-B98415/2020|EPI_ISL_675087|20 ···················· ···················· ····················· ···················· ·····T·············· ····················
U 2138 hCoV-19/England/MILK-B96C3D/2020|EPI_ISL_675088|20 ···················· ···················· ····················· ···················· ···················· ····················

| | | | | | | | |
|---|---|---|---|---|---|---|---|
| U | 2139 hCoV-19/England/MILK-B9708B/2020\|EPI_ISL_675089\|20 | ···················· | ···················· | ···················· | ···················· | ···················· | ···················· |
| U | 2140 hCoV-19/England/MILK-B973A0/2020\|EPI_ISL_675090\|20 | ···················· | ···················· | ···················· | ···················· | ······T············· | ···················· |
| U | 2141 hCoV-19/England/MILK-B962A4/2020\|EPI_ISL_675092\|20 | ···················· | ···················· | ···················· | ···················· | ···················· | ···················· |
| U | 2142 hCoV-19/England/MILK-B95A8F/2020\|EPI_ISL_675093\|20 | ···················· | ·A·················· | ···················· | ···················· | ···················· | ···A················ |
| U | 2143 hCoV-19/England/CAMC-B986B5/2020\|EPI_ISL_675094\|20 | ···················· | ·A·················· | ···················· | ···················· | ···················· | ···A················ |
| U | 2144 hCoV-19/England/MILK-B97391/2020\|EPI_ISL_675095\|20 | ···················· | ···················· | ···················· | ···················· | ···················· | ···················· |
| U | 2145 hCoV-19/England/MILK-B96286/2020\|EPI_ISL_675097\|20 | ···················· | ···················· | ···················· | ···················· | ···················· | ···················· |
| U | 2146 hCoV-19/England/MILK-B95E1A/2020\|EPI_ISL_675099\|20 | ···················· | ···················· | ···················· | ···················· | ···················· | ···················· |
| U | 2147 hCoV-19/England/MILK-B968B1/2020\|EPI_ISL_676300\|20 | ···················· | ···················· | ···················· | ···················· | ···················· | ···················· |
| U | 2148 hCoV-19/England/MILK-B9612F/2020\|EPI_ISL_676302\|20 | ···················· | ···················· | ···················· | ···················· | ···················· | ···················· |
| U | 2149 hCoV-19/England/MILK-B94419/2020\|EPI_ISL_676303\|20 | ···················· | ···················· | ···················· | ···················· | ···················· | ···················· |
| U | 2150 hCoV-19/England/MILK-B95DB3/2020\|EPI_ISL_676306\|20 | ···················· | ···················· | ···················· | ···················· | ···················· | ···················· |
| U | 2151 hCoV-19/England/MILK-B94710/2020\|EPI_ISL_676308\|20 | ···················· | ···················· | ···················· | ···················· | ···················· | ···················· |
| U | 2152 hCoV-19/England/MILK-B969AF/2020\|EPI_ISL_676310\|20 | ···················· | ···················· | ···················· | ···················· | ···················· | ···················· |
| U | 2153 hCoV-19/England/MILK-B94F27/2020\|EPI_ISL_676311\|20 | ···················· | ···················· | ···················· | ···················· | ···················· | ···················· |
| U | 2154 hCoV-19/Scotland/QEUH-B69495/2020\|EPI_ISL_676314\|2 | ···················· | ···················· | ···················· | ···················· | ···················· | ···················· |
| U | 2155 hCoV-19/Scotland/QEUH-B66E5E/2020\|EPI_ISL_676315\|2 | ···················· | ···················· | ···················· | ···················· | ···················· | ···················· |
| U | 2156 hCoV-19/Scotland/QEUH-B6943B/2020\|EPI_ISL_676316\|2 | ···················· | ···················· | ···················· | ···················· | ···················· | ···················· |
| U | 2157 hCoV-19/Scotland/QEUH-B694B3/2020\|EPI_ISL_676317\|2 | ···················· | ···················· | ···················· | ···················· | ···················· | ···················· |
| U | 2158 hCoV-19/Scotland/QEUH-B694FF/2020\|EPI_ISL_676319\|2 | ···················· | ···················· | ···················· | ···················· | ···················· | ···················· |
| U | 2159 hCoV-19/Scotland/QEUH-B66F4C/2020\|EPI_ISL_676320\|2 | ···················· | ···················· | ···················· | ···················· | ···················· | ···················· |
| U | 2160 hCoV-19/Scotland/QEUH-B694A4/2020\|EPI_ISL_676321\|2 | ···················· | ···················· | ···················· | ···················· | ···················· | ···················· |
| U | 2161 hCoV-19/Scotland/QEUH-B694E0/2020\|EPI_ISL_676322\|2 | ···················· | ···················· | ···················· | ···················· | ···················· | ···················· |
| U | 2162 hCoV-19/Scotland/QEUH-B66ED6/2020\|EPI_ISL_676323\|2 | ···················· | ···················· | ···················· | ···················· | ···················· | ···················· |
| U | 2163 hCoV-19/Scotland/QEUH-B69486/2020\|EPI_ISL_676326\|2 | ···················· | ···················· | ···················· | ···················· | ···················· | ···················· |
| U | 2164 hCoV-19/Scotland/QEUH-B681BD/2020\|EPI_ISL_676328\|2 | ···················· | ···················· | ···················· | ···················· | ···················· | ···················· |
| U | 2165 hCoV-19/Scotland/QEUH-B694C2/2020\|EPI_ISL_676329\|2 | ···················· | ···················· | ···················· | ···················· | ···················· | ···················· |
| U | 2166 hCoV-19/Scotland/QEUH-B66E4F/2020\|EPI_ISL_676330\|2 | ···················· | ···················· | ···················· | ···················· | ···················· | ···················· |
| U | 2167 hCoV-19/Scotland/QEUH-B6941D/2020\|EPI_ISL_676331\|2 | ···················· | ···················· | ···················· | ···················· | ···················· | ···················· |
| U | 2168 hCoV-19/Scotland/QEUH-B6944A/2020\|EPI_ISL_676332\|2 | ···················· | ···················· | ···················· | ···················· | ···················· | ···················· |
| U | 2169 hCoV-19/Scotland/QEUH-B69477/2020\|EPI_ISL_676333\|2 | ···················· | ···················· | ···················· | ···················· | ···················· | ···················· |
| U | 2170 hCoV-19/Scotland/QEUH-B69468/2020\|EPI_ISL_676335\|2 | ···················· | ···················· | ···················· | ···················· | ···················· | ···················· |
| U | 2171 hCoV-19/Scotland/QEUH-B675EF/2020\|EPI_ISL_676337\|2 | ···················· | ·A·················· | ···················· | ···················· | ···················· | ···················· |
| U | 2172 hCoV-19/Scotland/QEUH-B675A3/2020\|EPI_ISL_676338\|2 | ···················· | ···················· | ···················· | ···················· | ···················· | ···················· |
| U | 2173 hCoV-19/Scotland/QEUH-B66B39/2020\|EPI_ISL_676339\|2 | ···················· | ···················· | ···················· | ···················· | ···················· | ···················· |
| U | 2174 hCoV-19/Scotland/QEUH-B6A661/2020\|EPI_ISL_676340\|2 | ···················· | ···················· | ···················· | ···················· | ···················· | ···················· |
| U | 2175 hCoV-19/Scotland/QEUH-B6A29A/2020\|EPI_ISL_676341\|2 | ···················· | ···················· | ···················· | ···················· | ···················· | ···················· |
| U | 2176 hCoV-19/Scotland/QEUH-B6C580/2020\|EPI_ISL_676342\|2 | ···················· | ···················· | ···················· | ···················· | ···················· | ···················· |
| U | 2177 hCoV-19/Scotland/QEUH-B6C5F9/2020\|EPI_ISL_676343\|2 | ···················· | ···················· | ···················· | ···················· | ···················· | ···················· |
| U | 2178 hCoV-19/Scotland/QEUH-B6639B/2020\|EPI_ISL_676344\|2 | ···················· | ···················· | ···················· | ···················· | ···················· | ···················· |
| U | 2179 hCoV-19/Scotland/QEUH-B679C5/2020\|EPI_ISL_676345\|2 | ···················· | ···················· | ···················· | ···················· | ···················· | ···················· |
| U | 2180 hCoV-19/Scotland/QEUH-B66656/2020\|EPI_ISL_676346\|2 | ···················· | ···················· | ···················· | ···················· | ···················· | ···················· |
| U | 2181 hCoV-19/Scotland/QEUH-B67A95/2020\|EPI_ISL_676348\|2 | ···················· | ···················· | ···················· | ···················· | ···················· | ···················· |
| U | 2182 hCoV-19/Scotland/QEUH-B676A0/2020\|EPI_ISL_676349\|2 | ···················· | ···················· | ···················· | ···················· | ···················· | ···················· |
| U | 2183 hCoV-19/Scotland/QEUH-B66261/2020\|EPI_ISL_676350\|2 | ···················· | ···················· | ···················· | ·······C············ | ···················· | ···················· |
| U | 2184 hCoV-19/Scotland/QEUH-B81321/2020\|EPI_ISL_676353\|2 | ···················· | ···················· | ···················· | ···················· | ···················· | ···················· |
| U | 2185 hCoV-19/Scotland/QEUH-B7F46F/2020\|EPI_ISL_676354\|2 | ···················· | ···················· | ···················· | ···················· | ···················· | ···················· |
| U | 2186 hCoV-19/Scotland/QEUH-B7E390/2020\|EPI_ISL_676355\|2 | ···················· | ···················· | ···················· | ···················· | ···················· | ···················· |
| U | 2187 hCoV-19/Scotland/QEUH-B7FE8F/2020\|EPI_ISL_676356\|2 | ···················· | ···················· | ···················· | ···················· | ···················· | ···················· |
| U | 2188 hCoV-19/Scotland/QEUH-B7F441/2020\|EPI_ISL_676357\|2 | ···················· | ···················· | ···················· | ···················· | ···················· | ···················· |
| U | 2189 hCoV-19/Scotland/QEUH-B687AC/2020\|EPI_ISL_676359\|2 | ···················· | ···················· | ···················· | ···················· | ···················· | ···················· |
| U | 2190 hCoV-19/Scotland/QEUH-B69574/2020\|EPI_ISL_676363\|2 | ···················· | ···················· | ···················· | ···················· | ···················· | ···················· |
| U | 2191 hCoV-19/Scotland/QEUH-B68BBF/2020\|EPI_ISL_676364\|2 | ···················· | ···················· | ···················· | ···················· | ···················· | ···················· |
| U | 2192 hCoV-19/Scotland/QEUH-B68BEC/2020\|EPI_ISL_676365\|2 | ···················· | ···················· | ···················· | ···················· | ···················· | ···················· |
| U | 2193 hCoV-19/Scotland/QEUH-B69626/2020\|EPI_ISL_676366\|2 | ···················· | ···················· | ···················· | ···················· | ···················· | ···················· |
| U | 2194 hCoV-19/Scotland/QEUH-B68108/2020\|EPI_ISL_676368\|2 | ···················· | ···················· | ···················· | ···················· | ···················· | ···················· |
| U | 2195 hCoV-19/Scotland/QEUH-B69705/2020\|EPI_ISL_676369\|2 | ···················· | ···················· | ···················· | ···················· | ···················· | ···················· |
| U | 2196 hCoV-19/Scotland/QEUH-B69617/2020\|EPI_ISL_676380\|2 | ···················· | ···················· | ···················· | ···················· | ···················· | ···················· |
| U | 2197 hCoV-19/Scotland/QEUH-B68BA0/2020\|EPI_ISL_676381\|2 | ···················· | ···················· | ···················· | ···················· | ···················· | ···················· |
| U | 2198 hCoV-19/England/QEUH-B66067/2020\|EPI_ISL_676382\|20 | ···················· | ···················· | ···················· | ···················· | ···················· | ···················· |
| U | 2199 hCoV-19/Scotland/CAMC-B524EA/2020\|EPI_ISL_676383\|2 | ···················· | ···················· | ···················· | ···················· | ···················· | ···················· |
| U | 2200 hCoV-19/Scotland/QEUH-B403D3/2020\|EPI_ISL_676384\|2 | ···················· | ···················· | ···················· | ···················· | ···················· | ···················· |
| U | 2201 hCoV-19/Scotland/QEUH-B403B5/2020\|EPI_ISL_676385\|2 | ···················· | ···················· | ···················· | ···················· | ···················· | ···················· |
| U | 2202 hCoV-19/Scotland/QEUH-B40388/2020\|EPI_ISL_676386\|2 | ···················· | ···················· | ···················· | ···················· | ···················· | ···················· |
| U | 2203 hCoV-19/Scotland/QEUH-B4040D/2020\|EPI_ISL_676387\|2 | ···················· | ···················· | ···················· | ···················· | ···················· | ···················· |
| U | 2204 hCoV-19/Scotland/QEUH-B403F1/2020\|EPI_ISL_676388\|2 | ···················· | ···················· | ···················· | ···················· | ···················· | ···················· |
| U | 2205 hCoV-19/Scotland/QEUH-B4033D/2020\|EPI_ISL_676389\|2 | ···················· | ···················· | ···················· | ···················· | ···················· | ···················· |
| U | 2206 hCoV-19/Scotland/QEUH-B40036/2020\|EPI_ISL_676390\|2 | ···················· | ···················· | ···················· | ···················· | ···················· | ···················· |
| U | 2207 hCoV-19/Scotland/QEUH-B403C4/2020\|EPI_ISL_676391\|2 | ···················· | ···················· | ···················· | ···················· | ···················· | ···················· |
| U | 2208 hCoV-19/Scotland/QEUH-B4042B/2020\|EPI_ISL_676392\|2 | ···················· | ···················· | ···················· | ···················· | ···················· | ···················· |
| U | 2209 hCoV-19/Scotland/QEUH-B4035B/2020\|EPI_ISL_676393\|2 | ···················· | ···················· | ···················· | ···················· | ···················· | ···················· |
| U | 2210 hCoV-19/Scotland/QEUH-B402F4/2020\|EPI_ISL_676394\|2 | ···················· | ···················· | ···················· | ···················· | ···················· | ···················· |
| U | 2211 hCoV-19/Scotland/QEUH-B2EFB9/2020\|EPI_ISL_676395\|2 | ···················· | ···················· | ···················· | ···················· | ···················· | ···················· |
| U | 2212 hCoV-19/Scotland/QEUH-B2CCD2/2020\|EPI_ISL_676396\|2 | ···················· | ···················· | ···················· | ···················· | ···················· | ···················· |
| U | 2213 hCoV-19/Scotland/QEUH-B2CCE1/2020\|EPI_ISL_676397\|2 | ···················· | ···················· | ···················· | ···················· | ···················· | ···················· |
| U | 2214 hCoV-19/Scotland/QEUH-B2CC96/2020\|EPI_ISL_676398\|2 | ···················· | ···················· | ···················· | ···················· | ···················· | ···················· |
| U | 2215 hCoV-19/Scotland/QEUH-B2D6B5/2020\|EPI_ISL_676399\|2 | ···················· | ···················· | ···················· | ···················· | ···················· | ···················· |
| U | 2219 hCoV-19/USA/WI-UW-2224/2020\|EPI_ISL_677627\|2020-10 | ···················· | ···················· | ···················· | ···················· | ···················· | ···················· |
| U | 2221 hCoV-19/USA/WI-UW-2226/2020\|EPI_ISL_677629\|2020-10 | ···················· | ···················· | ···················· | ···················· | ···················· | ···················· |
| U | 2222 hCoV-19/USA/WI-UW-2227/2020\|EPI_ISL_677630\|2020-10 | ···················· | ···················· | ···················· | ···················· | ···················· | ···················· |

| | | | | | | | |
|---|---|---|---|---|---|---|---|
| U | 2223 hCoV-19/USA/WI-UW-2228/2020\|EPI_ISL_677631\|2020-10 | ···················· | ···················· | ···················· | ···················· | ···················· | ···················· |
| U | 2225 hCoV-19/USA/WI-UW-2230/2020\|EPI_ISL_677633\|2020-11 | ·····N·············· | ···················· | ···················· | ···················· | ···················· | ···················· |
| U | 2226 hCoV-19/Madagascar/VIRO-33698/2020\|EPI_ISL_677634\| | ···················· | ···················· | ···················· | ···················· | ···················· | ···················· |
| U | 2227 hCoV-19/Madagascar/VIRO-2722/2020\|EPI_ISL_677635\|2 | ···················· | ···················· | ···················· | ···················· | ···················· | ···················· |
| U | 2228 hCoV-19/Madagascar/VIRO-2728/2020\|EPI_ISL_677636\|2 | ···················· | ···················· | ···················· | ···················· | ···················· | ···················· |
| U | 2229 hCoV-19/USA/CO-CDPHE-2009021317/2020\|EPI_ISL_67763 | ···················· | ·A·················· | ···················· | ···················· | ···················· | ···················· |
| U | 2230 hCoV-19/USA/CO-CDPHE-2009021339/2020\|EPI_ISL_67763 | ···················· | ·A·················· | ···················· | ···················· | ···················· | ···················· |
| U | 2231 hCoV-19/USA/CO-CDPHE-2009042507/2020\|EPI_ISL_67764 | ···················· | ·A·················· | ···················· | ···················· | ···················· | ···················· |
| U | 2232 hCoV-19/USA/CO-CDPHE-2009042555/2020\|EPI_ISL_67764 | ···················· | ·A·················· | ···················· | ···················· | ···················· | ···················· |
| U | 2233 hCoV-19/USA/CO-CDPHE-2009090314/2020\|EPI_ISL_67764 | ···················· | ·A·················· | ···················· | ···················· | ···················· | ···················· |
| U | 2234 hCoV-19/USA/CO-CDPHE-2009090320/2020\|EPI_ISL_67764 | ···················· | ·A·················· | ···················· | ···················· | ···················· | ···················· |
| U | 2236 hCoV-19/USA/CO-CDPHE-2009090330/2020\|EPI_ISL_67764 | ···················· | ·A·················· | ···················· | ···················· | ···················· | ···················· |
| U | 2237 hCoV-19/USA/CO-CDPHE-2009163250/2020\|EPI_ISL_67764 | ···················· | ·A·················· | ···················· | ···················· | ···················· | ···················· |
| U | 2238 hCoV-19/USA/CO-CDPHE-2009163722/2020\|EPI_ISL_67765 | ···················· | ·A·················· | ···················· | ···················· | ··N················· | N··················· |
| U | 2239 hCoV-19/USA/CO-CDPHE-2009164004/2020\|EPI_ISL_67765 | ···················· | ·A·················· | ···················· | ···················· | ···················· | ···················· |
| U | 2240 hCoV-19/USA/CO-CDPHE-2009164036/2020\|EPI_ISL_67765 | ···················· | ·A·················· | ···················· | ···················· | ···················· | ···················· |
| U | 2241 hCoV-19/USA/WI-UW-1943/2020\|EPI_ISL_677658\|2020-10 | ···················· | ···················· | ···················· | ···················· | ···················· | ···················· |
| U | 2242 hCoV-19/USA/WI-UW-2013/2020\|EPI_ISL_677659\|2020-09 | ···················· | ···················· | ···················· | ···················· | ···················· | ···················· |
| U | 2244 hCoV-19/USA/NY-Wadsworth-46276-01/2020\|EPI_ISL_677 | ···················· | ···················· | ···················· | ···················· | ···················· | ···················· |
| U | 2245 hCoV-19/USA/NY-Wadsworth-81624-01/2020\|EPI_ISL_677 | ···················· | ····A··············· | ···················· | ···················· | ···················· | ···················· |
| U | 2246 hCoV-19/USA/NY-Wadsworth-10492-01/2020\|EPI_ISL_677 | ···················· | ···················· | ···················· | ···················· | ···················· | ···················· |
| U | 2247 hCoV-19/North Macedonia/883/2020\|EPI_ISL_677674\|20 | ···················· | ···················· | ···················· | ···················· | ···················· | ···················· |
| U | 2248 hCoV-19/North Macedonia/2052/2020\|EPI_ISL_677675\|2 | ···················· | ···················· | ···················· | ···················· | ···················· | ···················· |
| U | 2249 hCoV-19/North Macedonia/2227/2020\|EPI_ISL_677676\|2 | ···················· | ···················· | ···················· | ···················· | ···················· | ···················· |
| U | 2250 hCoV-19/North Macedonia/2339/2020\|EPI_ISL_677677\|2 | ···················· | ···················· | ···················· | ···················· | ···················· | ···················· |
| U | 2251 hCoV-19/USA/VAN3594/2020\|EPI_ISL_677678\|2020-04-17 | ···················· | ···················· | ···················· | ···················· | ···················· | ···················· |
| U | 2252 hCoV-19/USA/VAN4092/2020\|EPI_ISL_677679\|2020-04-25 | ···················· | ···················· | ···················· | ···················· | ···················· | ···················· |
| U | 2253 hCoV-19/USA/VAN3531/2020\|EPI_ISL_677680\|2020-04-13 | ···················· | ···················· | ···················· | ···················· | ···················· | ···················· |
| U | 2254 hCoV-19/USA/VAN3534/2020\|EPI_ISL_677681\|2020-04-11 | ···················· | ···················· | ···················· | ···················· | ···················· | ···················· |
| U | 2255 hCoV-19/USA/VAN3589/2020\|EPI_ISL_677682\|2020-04-17 | ···················· | ···················· | ···················· | ···················· | ···················· | ···················· |
| U | 2256 hCoV-19/USA/VAN3626/2020\|EPI_ISL_677683\|2020-04-20 | ···················· | ···················· | ···················· | ···················· | ···················· | ···················· |
| U | 2257 hCoV-19/USA/VAN3819/2020\|EPI_ISL_677684\|2020-04-30 | ···················· | ···················· | ···················· | ···················· | ···················· | ···················· |
| U | 2258 hCoV-19/USA/VAN4129/2020\|EPI_ISL_677685\|2020-05-06 | ···················· | ···················· | ···················· | ···················· | ···················· | ···················· |
| U | 2259 hCoV-19/USA/VAN4170/2020\|EPI_ISL_677686\|2020-05-07 | ···················· | ···················· | ···················· | ···················· | ···················· | ···················· |
| U | 2260 hCoV-19/USA/VAN4169/2020\|EPI_ISL_677687\|2020-05-11 | ···················· | ···················· | ···················· | ···················· | ···················· | ···················· |
| U | 2261 hCoV-19/USA/VAN4606/2020\|EPI_ISL_677688\|2020-05-11 | ···················· | ···················· | ···················· | ···················· | ···················· | ···················· |
| U | 2262 hCoV-19/USA/VAN3548/2020\|EPI_ISL_677689\|2020-04-15 | ···················· | ···················· | ···················· | ···················· | ···················· | ···················· |
| U | 2263 hCoV-19/USA/VAN5434/2020\|EPI_ISL_677699\|2020-09-16 | ···················· | ···················· | ···················· | ···················· | ···················· | ···················· |
| U | 2264 hCoV-19/England/ALDP-B748E6/2020\|EPI_ISL_678956\|20 | ···················· | ···················· | ···················· | ···················· | ···················· | ···················· |
| U | 2265 hCoV-19/England/ALDP-B748F5/2020\|EPI_ISL_678957\|20 | ···················· | ···················· | ···················· | ···················· | ···················· | ···················· |
| U | 2266 hCoV-19/England/ALDP-B74A4A/2020\|EPI_ISL_678961\|20 | ···················· | ···················· | ···················· | ···················· | ···················· | ···················· |
| U | 2267 hCoV-19/England/ALDP-B74A59/2020\|EPI_ISL_678962\|20 | ···················· | ···················· | ···················· | ···················· | ···················· | ···················· |
| U | 2268 hCoV-19/England/ALDP-B74A68/2020\|EPI_ISL_678963\|20 | ···················· | ···················· | ···················· | ···················· | ···················· | ···················· |
| U | 2269 hCoV-19/England/ALDP-B74A86/2020\|EPI_ISL_678965\|20 | ···················· | ···················· | ···················· | ···················· | ···················· | ···················· |
| U | 2270 hCoV-19/England/ALDP-B74A95/2020\|EPI_ISL_678966\|20 | ···················· | ···················· | ···················· | ···················· | ···················· | ···················· |
| U | 2271 hCoV-19/England/ALDP-B74AA4/2020\|EPI_ISL_678967\|20 | ···················· | ···················· | ···················· | ···················· | ···················· | ···················· |
| U | 2272 hCoV-19/England/ALDP-B74AC2/2020\|EPI_ISL_678969\|20 | ···················· | ···················· | ···················· | ···················· | ···················· | ···················· |
| U | 2273 hCoV-19/England/ALDP-B74AD1/2020\|EPI_ISL_678970\|20 | ···················· | ···················· | ···················· | ···················· | ···················· | ···················· |
| U | 2274 hCoV-19/England/ALDP-B74AE0/2020\|EPI_ISL_678971\|20 | ···················· | ···················· | ···················· | ···················· | ···················· | ···················· |
| U | 2275 hCoV-19/England/ALDP-B74B0B/2020\|EPI_ISL_678972\|20 | ···················· | ···················· | ···················· | ···················· | ···················· | ···················· |
| U | 2276 hCoV-19/England/ALDP-B74B1A/2020\|EPI_ISL_678973\|20 | ···················· | ···················· | ···················· | ···················· | ···················· | ···················· |
| U | 2277 hCoV-19/England/ALDP-B74B38/2020\|EPI_ISL_678975\|20 | ···················· | ···················· | ···················· | ···················· | ···················· | ···················· |
| U | 2278 hCoV-19/England/ALDP-B74B56/2020\|EPI_ISL_678976\|20 | ···················· | ···················· | ···················· | ···················· | ······T············· | ···················· |
| U | 2279 hCoV-19/England/ALDP-B74B74/2020\|EPI_ISL_678978\|20 | ···················· | ···················· | ···················· | ···················· | ······T············· | ···················· |
| U | 2280 hCoV-19/England/ALDP-B74B92/2020\|EPI_ISL_678980\|20 | ···················· | ···················· | ···················· | ···················· | ···················· | ···················· |
| U | 2281 hCoV-19/England/ALDP-B74BB0/2020\|EPI_ISL_678982\|20 | ···················· | ···················· | ···················· | ···················· | ···················· | ···················· |
| U | 2282 hCoV-19/England/ALDP-B74BED/2020\|EPI_ISL_678985\|20 | ···················· | ···················· | ···················· | ···················· | ···················· | ···················· |
| U | 2283 hCoV-19/England/ALDP-B74BFC/2020\|EPI_ISL_678986\|20 | ···················· | ···················· | ···················· | ···················· | ···················· | ···················· |
| U | 2284 hCoV-19/England/ALDP-B74C08/2020\|EPI_ISL_678987\|20 | ···················· | ···················· | ···················· | ···················· | ······T············· | ···················· |
| U | 2285 hCoV-19/England/ALDP-B74C17/2020\|EPI_ISL_678988\|20 | ···················· | ···················· | ···················· | ···················· | ···················· | ···················· |
| U | 2286 hCoV-19/England/ALDP-B74C26/2020\|EPI_ISL_678989\|20 | ···················· | ···················· | ···················· | ···················· | ···················· | ···················· |
| U | 2287 hCoV-19/England/ALDP-B74C35/2020\|EPI_ISL_678990\|20 | ···················· | ···················· | ···················· | ···················· | ······T············· | ···················· |
| U | 2288 hCoV-19/England/ALDP-B74C44/2020\|EPI_ISL_678991\|20 | ···················· | ···················· | ···················· | ···················· | ···················· | ···················· |
| U | 2289 hCoV-19/England/ALDP-B74DF6/2020\|EPI_ISL_678992\|20 | ···················· | ···················· | ···················· | ···················· | ···················· | ···················· |
| U | 2290 hCoV-19/England/ALDP-B74E02/2020\|EPI_ISL_678993\|20 | ···················· | ···················· | ···················· | ···················· | ···················· | ···················· |
| U | 2291 hCoV-19/England/ALDP-B74E11/2020\|EPI_ISL_678994\|20 | ···················· | ···················· | ···················· | ···················· | ···················· | ···················· |
| U | 2292 hCoV-19/England/ALDP-B74E7B/2020\|EPI_ISL_678998\|20 | ···················· | ···················· | ···················· | ···················· | ···················· | ···················· |
| U | 2293 hCoV-19/USA/MI-UM-10036686888/2020\|EPI_ISL_682120\| | ···················· | ···················· | ···················· | ···················· | ···················· | ···················· |
| U | 2294 hCoV-19/USA/MI-UM-10036711590/2020\|EPI_ISL_682121\| | ···················· | ······A············· | ···················· | ···················· | ···················· | ···················· |
| U | 2295 hCoV-19/USA/MI-UM-10036704076/2020\|EPI_ISL_682123\| | ···················· | ···················· | ···················· | ···················· | ···················· | ···················· |
| U | 2296 hCoV-19/USA/MI-UM-10036735869/2020\|EPI_ISL_682124\| | ···················· | ···················· | ···················· | ···················· | ···················· | ···················· |
| U | 2297 hCoV-19/USA/MI-UM-10036692268/2020\|EPI_ISL_682126\| | ···················· | ······A············· | ···················· | ···················· | ···················· | ···················· |
| U | 2298 hCoV-19/USA/MI-UM-10036711844/2020\|EPI_ISL_682127\| | ···················· | ······A············· | ···················· | ···················· | ···················· | ···················· |
| U | 2299 hCoV-19/USA/MI-UM-10036738392/2020\|EPI_ISL_682129\| | ···················· | ·············A······ | ···················· | ···················· | ···················· | ···················· |
| U | 2300 hCoV-19/USA/MI-UM-10036712614/2020\|EPI_ISL_682131\| | ···················· | ···················· | ···················· | ···················· | ···················· | ···················· |
| U | 2301 hCoV-19/USA/MI-UM-10036702239/2020\|EPI_ISL_682132\| | ···················· | ···················· | ···················· | ···················· | ···················· | ···················· |
| U | 2303 hCoV-19/Armenia/ChVir8660/2020\|EPI_ISL_683450\|2020 | ···················· | ···················· | ···················· | ···················· | ···················· | ···················· |
| U | 2304 hCoV-19/Armenia/ChVir8668/2020\|EPI_ISL_683451\|2020 | ···················· | ···················· | ···················· | ···················· | ···················· | ···················· |
| U | 2305 hCoV-19/Brazil/SP-L28-CD686/2020\|EPI_ISL_672748\|20 | ···················· | ···················· | ···················· | ···················· | ···················· | ···················· |
| U | 2306 hCoV-19/England/QEUH-BA1279/2020\|EPI_ISL_672754\|20 | ···················· | ···················· | ···················· | ···················· | ···················· | ···················· |

| | | | | | | | |
|---|---|---|---|---|---|---|---|
| U | 2307 hCoV-19/England/QEUH-BA11E5/2020\|EPI_ISL_672758\|20 | ···················· | ···················· | ····················· | ···················· | ···················· | ···················· |
| U | 2309 hCoV-19/England/QEUH-BA2B9B/2020\|EPI_ISL_672760\|20 | ···················· | ···················· | ····················· | ···················· | ···················· | ···················· |
| U | 2310 hCoV-19/England/QEUH-BA126A/2020\|EPI_ISL_672761\|20 | ···················· | ···················· | ····················· | ···················· | ···················· | ···················· |
| U | 2311 hCoV-19/England/QEUH-BA1E66/2020\|EPI_ISL_672764\|20 | ···················· | ···················· | ····················· | ···················· | ···················· | ···················· |
| U | 2312 hCoV-19/England/QEUH-BA2EDE/2020\|EPI_ISL_672765\|20 | ···················· | ···················· | ····················· | ···················· | ···················· | ···················· |
| U | 2313 hCoV-19/England/QEUH-BA11F4/2020\|EPI_ISL_672767\|20 | ···················· | ···················· | ····················· | ···················· | ···················· | ···················· |
| U | 2314 hCoV-19/England/QEUH-BA26B8/2020\|EPI_ISL_672768\|20 | ···················· | ···················· | ····················· | ···················· | ···················· | ···················· |
| U | 2315 hCoV-19/England/QEUH-BA0E2B/2020\|EPI_ISL_672769\|20 | ···················· | ···················· | ····················· | ···················· | ···················· | ···················· |
| U | 2316 hCoV-19/England/QEUH-BA3426/2020\|EPI_ISL_672770\|20 | ···················· | ···················· | ····················· | ···················· | ···················· | ···················· |
| U | 2318 hCoV-19/England/QEUH-BA2937/2020\|EPI_ISL_672774\|20 | ···················· | ···················· | ····················· | ···················· | ···················· | ···················· |
| U | 2319 hCoV-19/England/QEUH-BA1288/2020\|EPI_ISL_672775\|20 | ···················· | ···················· | ····················· | ···················· | ···················· | ···················· |
| U | 2320 hCoV-19/England/QEUH-BA11A9/2020\|EPI_ISL_672777\|20 | ···················· | ·A·················· | ····················· | ···················· | ···················· | ··A················· |
| U | 2321 hCoV-19/England/QEUH-BA30E6/2020\|EPI_ISL_672778\|20 | ···················· | ···················· | ····················· | ···················· | ···················· | ···················· |
| U | 2322 hCoV-19/England/QEUH-BA2EA1/2020\|EPI_ISL_672780\|20 | ···················· | ···················· | ····················· | ···················· | ···················· | ···················· |
| U | 2323 hCoV-19/England/QEUH-BA1394/2020\|EPI_ISL_672781\|20 | ···················· | ···················· | ····················· | ···················· | ···················· | ···················· |
| U | 2324 hCoV-19/England/QEUH-BA2A61/2020\|EPI_ISL_672782\|20 | ···················· | ···················· | ····················· | ···················· | ···················· | ···················· |
| U | 2325 hCoV-19/England/QEUH-BA316B/2020\|EPI_ISL_672786\|20 | ···················· | ···················· | ····················· | ···················· | ···················· | ···················· |
| U | 2326 hCoV-19/England/QEUH-BA13EF/2020\|EPI_ISL_672787\|20 | ···················· | ···················· | ····················· | ···················· | ···················· | ···················· |
| U | 2327 hCoV-19/England/QEUH-BA1F27/2020\|EPI_ISL_672788\|20 | ···················· | ···················· | ····················· | ···················· | ···················· | ···················· |
| U | 2328 hCoV-19/England/QEUH-BA122E/2020\|EPI_ISL_672789\|20 | ···················· | ···················· | ····················· | ···················· | ···················· | ···················· |
| U | 2329 hCoV-19/England/QEUH-BA31B6/2020\|EPI_ISL_672790\|20 | ···················· | ···················· | ····················· | ···················· | ···················· | ···················· |
| U | 2330 hCoV-19/England/QEUH-BA2A07/2020\|EPI_ISL_672793\|20 | ···················· | ···················· | ····················· | ···················· | ···················· | ···················· |
| U | 2331 hCoV-19/England/MILK-B96778/2020\|EPI_ISL_676210\|20 | ···················· | ···················· | ····················· | ···················· | ···················· | ···················· |
| U | 2332 hCoV-19/England/MILK-B94DA5/2020\|EPI_ISL_676216\|20 | ···················· | ·A·················· | ····················· | ···················· | ···················· | ··A················· |
| U | 2333 hCoV-19/England/MILK-B96B6D/2020\|EPI_ISL_676217\|20 | ···················· | ···················· | ····················· | ···················· | ···················· | ···················· |
| U | 2334 hCoV-19/England/MILK-B95D86/2020\|EPI_ISL_676219\|20 | ···················· | ···················· | ····················· | ···················· | ···················· | ···················· |
| U | 2335 hCoV-19/England/MILK-B967C3/2020\|EPI_ISL_676221\|20 | ···················· | ···················· | ····················· | ···················· | ···················· | ···················· |
| U | 2336 hCoV-19/England/MILK-B96347/2020\|EPI_ISL_676223\|20 | ···················· | ···················· | ····················· | ···················· | ···················· | ···················· |
| U | 2337 hCoV-19/England/MILK-B94E48/2020\|EPI_ISL_676226\|20 | ···················· | ···················· | ····················· | ···················· | ···················· | ···················· |
| U | 2338 hCoV-19/England/MILK-B948A4/2020\|EPI_ISL_676227\|20 | ···················· | ···················· | ····················· | ···················· | ···················· | ···················· |
| U | 2339 hCoV-19/England/MILK-B943D0/2020\|EPI_ISL_676228\|20 | ···················· | ···················· | ····················· | ···················· | ···················· | ···················· |
| U | 2340 hCoV-19/England/MILK-B93DA6/2020\|EPI_ISL_676229\|20 | ···················· | ···················· | ····················· | ···················· | ·····T·············· | ···················· |
| U | 2341 hCoV-19/England/MILK-B95D1D/2020\|EPI_ISL_676231\|20 | ···················· | ···················· | ····················· | ···················· | ···················· | ···················· |
| U | 2342 hCoV-19/England/MILK-B93D5B/2020\|EPI_ISL_676232\|20 | ·T·················· | ···················· | ····················· | ···················· | ···················· | ···················· |
| U | 2343 hCoV-19/England/MILK-B96769/2020\|EPI_ISL_676233\|20 | ···················· | ···················· | ····················· | ···················· | ···················· | ···················· |
| U | 2344 hCoV-19/England/MILK-B96839/2020\|EPI_ISL_676234\|20 | ···················· | ···················· | ····················· | ···················· | ···················· | ···················· |
| U | 2345 hCoV-19/England/MILK-B9608C/2020\|EPI_ISL_676235\|20 | ···················· | ···················· | ····················· | ···················· | ···················· | ···················· |
| U | 2346 hCoV-19/England/MILK-B94E0C/2020\|EPI_ISL_676236\|20 | ···················· | ···················· | ····················· | ···················· | ···················· | ···················· |
| U | 2347 hCoV-19/England/MILK-B963ED/2020\|EPI_ISL_676237\|20 | ···················· | ···················· | ····················· | ···················· | ···················· | ···················· |
| U | 2348 hCoV-19/England/MILK-B94464/2020\|EPI_ISL_676238\|20 | ···················· | ···················· | ····················· | ···················· | ···················· | ···················· |
| U | 2349 hCoV-19/England/MILK-B96426/2020\|EPI_ISL_676239\|20 | ···················· | ···················· | ····················· | ···················· | ···················· | ···················· |
| U | 2350 hCoV-19/England/MILK-B94A44/2020\|EPI_ISL_676240\|20 | ···················· | ···················· | ····················· | ···················· | ···················· | ···················· |
| U | 2351 hCoV-19/England/MILK-B94103/2020\|EPI_ISL_676241\|20 | ···················· | ·A·················· | ····················· | ···················· | ···················· | ··A················· |
| U | 2352 hCoV-19/England/MILK-B94A35/2020\|EPI_ISL_676242\|20 | ···················· | ···················· | ····················· | ···················· | ···················· | ···················· |
| U | 2353 hCoV-19/England/MILK-B965F6/2020\|EPI_ISL_676243\|20 | ···················· | ···················· | ····················· | ···················· | ···················· | ···················· |
| U | 2354 hCoV-19/England/MILK-B948C2/2020\|EPI_ISL_676244\|20 | ···················· | ···················· | ····················· | ···················· | ···················· | ···················· |
| U | 2355 hCoV-19/England/MILK-B95F26/2020\|EPI_ISL_676245\|20 | ···················· | ···················· | ····················· | ···················· | ···················· | ···················· |
| U | 2356 hCoV-19/England/MILK-B944EC/2020\|EPI_ISL_676246\|20 | ···················· | ···················· | ····················· | ···················· | ···················· | ···················· |
| U | 2357 hCoV-19/England/MILK-B96A42/2020\|EPI_ISL_676247\|20 | ···················· | ···················· | ····················· | ···················· | ···················· | ···················· |
| U | 2358 hCoV-19/England/MILK-B968DF/2020\|EPI_ISL_676248\|20 | ···················· | ···················· | ····················· | ···················· | ···················· | ···················· |
| U | 2359 hCoV-19/England/MILK-B962D1/2020\|EPI_ISL_676249\|20 | ···················· | ·A·················· | ····················· | ···················· | ···················· | ··A················· |
| U | 2360 hCoV-19/England/MILK-B95C01/2020\|EPI_ISL_676250\|20 | ···················· | ···················· | ····················· | ···················· | ···················· | ···················· |
| U | 2361 hCoV-19/England/MILK-B94F45/2020\|EPI_ISL_676251\|20 | ···················· | ···················· | ····················· | ···················· | ·····T·············· | ···················· |
| U | 2362 hCoV-19/England/MILK-B9665D/2020\|EPI_ISL_676252\|20 | ···················· | ···················· | ····················· | ···················· | ···················· | ···················· |
| U | 2363 hCoV-19/England/MILK-B94EFD/2020\|EPI_ISL_676253\|20 | ···················· | ···················· | ····················· | ···················· | ···················· | ···················· |
| U | 2364 hCoV-19/England/MILK-B96918/2020\|EPI_ISL_676254\|20 | ···················· | ···················· | ····················· | ···················· | ···················· | ···················· |
| U | 2365 hCoV-19/England/MILK-B944CE/2020\|EPI_ISL_676255\|20 | ···················· | ···················· | ····················· | ···················· | ···················· | ···················· |
| U | 2366 hCoV-19/England/MILK-B965C9/2020\|EPI_ISL_676256\|20 | ···················· | ···················· | ····················· | ···················· | ···················· | ···················· |
| U | 2367 hCoV-19/England/MILK-B93BCA/2020\|EPI_ISL_676257\|20 | ···················· | ···················· | ····················· | ···················· | ···················· | ···················· |
| U | 2368 hCoV-19/England/MILK-B949B0/2020\|EPI_ISL_676258\|20 | ···················· | ···················· | ····················· | ···················· | ···················· | ···················· |
| U | 2369 hCoV-19/USA/WI-UW-2136/2020\|EPI_ISL_677540\|2020-10 | ···················· | ···················· | ····················· | ···················· | ···················· | ···················· |
| U | 2370 hCoV-19/USA/WI-UW-2143/2020\|EPI_ISL_677547\|2020-10 | ···················· | ···················· | ····················· | ···················· | ···················· | ···················· |
| U | 2383 hCoV-19/USA/WI-UW-2197/2020\|EPI_ISL_677600\|2020-10 | ···················· | ···················· | ····················· | ···················· | ···················· | ···················· |
| U | 2384 hCoV-19/USA/WI-UW-2198/2020\|EPI_ISL_677601\|2020-10 | ···················· | ···················· | ····················· | ···················· | ···················· | ···················· |
| U | 2385 hCoV-19/USA/WI-UW-2199/2020\|EPI_ISL_677602\|2020-10 | ···················· | ···················· | ····················· | ···················· | ···················· | ···················· |
| U | 2388 hCoV-19/USA/WI-UW-2202/2020\|EPI_ISL_677605\|2020-10 | ···················· | ···················· | ····················· | ···················· | ···················· | ···················· |
| U | 2389 hCoV-19/USA/WI-UW-2203/2020\|EPI_ISL_677606\|2020-10 | ···················· | ···················· | ····················· | ···················· | ···················· | ···················· |
| U | 2390 hCoV-19/USA/WI-UW-2204/2020\|EPI_ISL_677607\|2020-10 | ···················· | ···················· | ····················· | ···················· | ···················· | ···················· |
| U | 2391 hCoV-19/USA/WI-UW-2205/2020\|EPI_ISL_677608\|2020-10 | ···················· | ···················· | ····················· | ···················· | ···················· | ···················· |
| U | 2392 hCoV-19/USA/WI-UW-2206/2020\|EPI_ISL_677609\|2020-10 | ···················· | ···················· | ····················· | ···················· | ···················· | ···················· |
| U | 2393 hCoV-19/USA/WI-UW-2207/2020\|EPI_ISL_677610\|2020-10 | ···················· | ···················· | ····················· | ···················· | ···················· | ···················· |
| U | 2394 hCoV-19/USA/WI-UW-2209/2020\|EPI_ISL_677612\|2020-10 | ···················· | ···················· | ····················· | ···················· | ···················· | ···················· |
| U | 2395 hCoV-19/USA/WI-UW-2210/2020\|EPI_ISL_677613\|2020-10 | ···················· | ···················· | ····················· | ···················· | ···················· | ···················· |
| U | 2396 hCoV-19/USA/WI-UW-2211/2020\|EPI_ISL_677614\|2020-10 | ···················· | ·N·················· | ····················· | ···················· | ···················· | ···················· |
| U | 2397 hCoV-19/USA/WI-UW-2212/2020\|EPI_ISL_677615\|2020-10 | ···················· | ···················· | ····················· | ···················· | ···················· | ···················· |
| U | 2398 hCoV-19/USA/WI-UW-2213/2020\|EPI_ISL_677616\|2020-10 | ···················· | ···················· | ····················· | ···················· | ···················· | ···················· |
| U | 2399 hCoV-19/USA/WI-UW-2214/2020\|EPI_ISL_677617\|2020-10 | ···················· | ···················· | ····················· | ···················· | ···················· | ···················· |
| U | 2401 hCoV-19/USA/WI-UW-2216/2020\|EPI_ISL_677619\|2020-09 | ···················· | ···················· | ····················· | ···················· | ···················· | ···················· |
| U | 2402 hCoV-19/USA/WI-UW-2217/2020\|EPI_ISL_677620\|2020-09 | ···················· | ···················· | ····················· | ···················· | ···················· | ···················· |
| U | 2403 hCoV-19/USA/WI-UW-2218/2020\|EPI_ISL_677621\|2020-09 | ···················· | ···················· | ····················· | ···················· | ···················· | ···················· |

| | | | | | | | | |
|---|---|---|---|---|---|---|---|---|
| U | 2404 | hCoV-19/USA/WI-UW-2219/2020\|EPI_ISL_677622\|2020-09 | ···················· | ···················· | ····················· | ···················· | ···················· | ···················· |
| U | 2405 | hCoV-19/USA/WI-UW-2220/2020\|EPI_ISL_677623\|2020-09 | ···················· | ···················· | ····················· | ···················· | ···················· | ···················· |
| U | 2406 | hCoV-19/England/ALDP-B5D1E8/2020\|EPI_ISL_678870\|20 | ···················· | ···················· | ····················· | ···················· | ······T············· | ···················· |
| U | 2407 | hCoV-19/England/ALDP-B5D203/2020\|EPI_ISL_678872\|20 | ···················· | ···················· | ····················· | ···················· | ···················· | ···················· |
| U | 2408 | hCoV-19/England/ALDP-B5D230/2020\|EPI_ISL_678875\|20 | ···················· | ···················· | ····················· | ···················· | ···················· | ···················· |
| U | 2409 | hCoV-19/England/ALDP-B5D28B/2020\|EPI_ISL_678880\|20 | ···················· | ···················· | ····················· | ···················· | ······T············· | ···················· |
| U | 2410 | hCoV-19/England/ALDP-B5D29A/2020\|EPI_ISL_678881\|20 | ···················· | ···················· | ····················· | ···················· | ······T············· | ···················· |
| U | 2411 | hCoV-19/England/ALDP-B5D2A9/2020\|EPI_ISL_678882\|20 | ···················· | ···················· | ····················· | ···················· | ···················· | ···················· |
| U | 2412 | hCoV-19/England/ALDP-B5D2D6/2020\|EPI_ISL_678884\|20 | ···················· | ···················· | ····················· | ···················· | ···················· | ···················· |
| U | 2413 | hCoV-19/England/ALDP-B5D2F4/2020\|EPI_ISL_678885\|20 | ···················· | ·A·················· | ····················· | ···················· | ···················· | ···A················ |
| U | 2414 | hCoV-19/England/ALDP-B5D31F/2020\|EPI_ISL_678887\|20 | ···················· | ·A·················· | ····················· | ···················· | ···················· | ···················· |
| U | 2415 | hCoV-19/England/ALDP-B5D33D/2020\|EPI_ISL_678889\|20 | ···················· | ···················· | ····················· | ···················· | ···················· | ···················· |
| U | 2416 | hCoV-19/England/ALDP-B5D35B/2020\|EPI_ISL_678890\|20 | ···················· | ···················· | ····················· | ···················· | ···················· | ···················· |
| U | 2417 | hCoV-19/England/ALDP-B5D388/2020\|EPI_ISL_678892\|20 | ···················· | ···················· | ····················· | ···················· | ···················· | ···················· |
| U | 2418 | hCoV-19/England/ALDP-B5D3A6/2020\|EPI_ISL_678893\|20 | ···················· | ···················· | ····················· | ···················· | ···················· | ···················· |
| U | 2419 | hCoV-19/England/ALDP-B5D3C4/2020\|EPI_ISL_678894\|20 | ···················· | ···················· | ····················· | ···················· | ···················· | ···················· |
| U | 2420 | hCoV-19/England/ALDP-B5D3E2/2020\|EPI_ISL_678895\|20 | ···················· | ···················· | ····················· | ···················· | ···················· | ···················· |
| U | 2421 | hCoV-19/England/ALDP-B5D3F1/2020\|EPI_ISL_678896\|20 | ···················· | ···················· | ····················· | ···················· | ···················· | ···················· |
| U | 2422 | hCoV-19/England/ALDP-B5D42B/2020\|EPI_ISL_678898\|20 | ···················· | ···················· | ····················· | ···················· | ···················· | ···················· |
| U | 2423 | hCoV-19/England/ALDP-B5D449/2020\|EPI_ISL_678899\|20 | ···················· | ···················· | ····················· | ···················· | ···················· | ···················· |
| U | 2424 | hCoV-19/England/ALDP-B5D4B2/2020\|EPI_ISL_678901\|20 | ···················· | ···················· | ····················· | ···················· | ···················· | ···················· |
| U | 2425 | hCoV-19/England/ALDP-B5D4C1/2020\|EPI_ISL_678902\|20 | ···················· | ·A·················· | ····················· | ···················· | ···················· | ···A················ |
| U | 2426 | hCoV-19/England/ALDP-B5D4D0/2020\|EPI_ISL_678903\|20 | ···················· | ···················· | ····················· | ···················· | ···················· | ···················· |
| U | 2427 | hCoV-19/England/ALDP-B5D4FE/2020\|EPI_ISL_678905\|20 | ···················· | ···················· | ····················· | ···················· | ···················· | ···················· |
| U | 2428 | hCoV-19/England/ALDP-B5D50A/2020\|EPI_ISL_678906\|20 | ···················· | ···················· | ····················· | ···················· | ···················· | ···················· |
| U | 2429 | hCoV-19/England/ALDP-B5D519/2020\|EPI_ISL_678907\|20 | ···················· | ···················· | ····················· | ···················· | ···················· | ···················· |
| U | 2430 | hCoV-19/England/ALDP-B5D537/2020\|EPI_ISL_678909\|20 | ···················· | ···················· | ····················· | ···················· | ···················· | ···················· |
| U | 2431 | hCoV-19/England/ALDP-B5D546/2020\|EPI_ISL_678910\|20 | ···················· | ···················· | ····················· | ···················· | ···················· | ···················· |
| U | 2432 | hCoV-19/England/ALDP-B5D555/2020\|EPI_ISL_678911\|20 | ···················· | ···················· | ····················· | ···················· | ···················· | ···················· |
| U | 2433 | hCoV-19/England/ALDP-B5D564/2020\|EPI_ISL_678912\|20 | ···················· | ···················· | ····················· | ···················· | ···················· | ···················· |
| U | 2434 | hCoV-19/England/ALDP-B5D573/2020\|EPI_ISL_678913\|20 | ···················· | ···················· | ····················· | ···················· | ···················· | ···················· |
| U | 2435 | hCoV-19/England/ALDP-B5D591/2020\|EPI_ISL_678915\|20 | ···················· | ···················· | ····················· | ···················· | ···················· | ···················· |
| U | 2437 | hCoV-19/England/ALDP-B741FA/2020\|EPI_ISL_678919\|20 | ···················· | ···················· | ····················· | ···················· | ···················· | ···················· |
| U | 2438 | hCoV-19/England/ALDP-B74479/2020\|EPI_ISL_678920\|20 | ···················· | ···················· | ····················· | ···················· | ···················· | ···················· |
| U | 2439 | hCoV-19/England/ALDP-B744C4/2020\|EPI_ISL_678921\|20 | ···················· | ···················· | ····················· | ···················· | ···················· | ···················· |
| U | 2440 | hCoV-19/England/ALDP-B7453A/2020\|EPI_ISL_678923\|20 | ···················· | ···················· | ····················· | ···················· | ···················· | ···················· |
| U | 2441 | hCoV-19/England/ALDP-B74558/2020\|EPI_ISL_678925\|20 | ···················· | ···················· | ····················· | ···················· | ···················· | ···················· |
| U | 2442 | hCoV-19/England/ALDP-B74576/2020\|EPI_ISL_678927\|20 | ···················· | ···················· | ····················· | ···················· | ···················· | ···················· |
| U | 2443 | hCoV-19/England/ALDP-B74585/2020\|EPI_ISL_678928\|20 | ···················· | ···················· | ····················· | ···················· | ···················· | ···················· |
| U | 2444 | hCoV-19/England/ALDP-B74594/2020\|EPI_ISL_678929\|20 | ···················· | ···················· | ····················· | ···················· | ···················· | ···················· |
| U | 2445 | hCoV-19/England/ALDP-B745A3/2020\|EPI_ISL_678930\|20 | ···················· | ···················· | ····················· | ···················· | ···················· | ···················· |
| U | 2446 | hCoV-19/England/ALDP-B745C1/2020\|EPI_ISL_678932\|20 | ···················· | ···················· | ····················· | ···················· | ···················· | ···················· |
| U | 2447 | hCoV-19/England/ALDP-B745D0/2020\|EPI_ISL_678933\|20 | ···················· | ···················· | ····················· | ···················· | ···················· | ···················· |
| U | 2448 | hCoV-19/England/ALDP-B745EF/2020\|EPI_ISL_678934\|20 | ···················· | ···················· | ····················· | ···················· | ···················· | ···················· |
| U | 2449 | hCoV-19/England/ALDP-B7460A/2020\|EPI_ISL_678936\|20 | ···················· | ···················· | ····················· | ···················· | ···················· | ···················· |
| U | 2450 | hCoV-19/England/ALDP-B74619/2020\|EPI_ISL_678937\|20 | ···················· | ···················· | ····················· | ···················· | ···················· | ···················· |
| U | 2451 | hCoV-19/England/ALDP-B74628/2020\|EPI_ISL_678938\|20 | ···················· | ···················· | ····················· | ···················· | ···················· | ···················· |
| U | 2452 | hCoV-19/England/ALDP-B747BC/2020\|EPI_ISL_678939\|20 | ···················· | ···················· | ····················· | ···················· | ······T············· | ···················· |
| U | 2453 | hCoV-19/England/ALDP-B747DA/2020\|EPI_ISL_678941\|20 | ···················· | ···················· | ····················· | ···················· | ···················· | ···················· |
| U | 2454 | hCoV-19/England/ALDP-B747F8/2020\|EPI_ISL_678942\|20 | ···················· | ···················· | ····················· | ···················· | ···················· | ···················· |
| U | 2455 | hCoV-19/England/ALDP-B74804/2020\|EPI_ISL_678943\|20 | ···················· | ···················· | ····················· | ···················· | ···················· | ···················· |
| U | 2456 | hCoV-19/England/ALDP-B74813/2020\|EPI_ISL_678944\|20 | ···················· | ···················· | ····················· | ···················· | ···················· | ···················· |
| U | 2457 | hCoV-19/England/ALDP-B74822/2020\|EPI_ISL_678945\|20 | ···················· | ···················· | ····················· | ···················· | ···················· | ···················· |
| U | 2458 | hCoV-19/England/ALDP-B74831/2020\|EPI_ISL_678946\|20 | ···················· | ···················· | ····················· | ···················· | ···················· | ···················· |
| U | 2459 | hCoV-19/England/ALDP-B74840/2020\|EPI_ISL_678947\|20 | ···················· | ···················· | ····················· | ···················· | ···················· | ···················· |
| U | 2460 | hCoV-19/England/ALDP-B7486E/2020\|EPI_ISL_678949\|20 | ···················· | ···················· | ····················· | ···················· | ···················· | ···················· |
| U | 2461 | hCoV-19/England/ALDP-B7489B/2020\|EPI_ISL_678951\|20 | ···················· | ···················· | ····················· | ···················· | ······T············· | ···················· |
| U | 2462 | hCoV-19/England/ALDP-B748AA/2020\|EPI_ISL_678952\|20 | ···················· | ···················· | ····················· | ···················· | ···················· | ···················· |
| U | 2463 | hCoV-19/England/ALDP-B748C8/2020\|EPI_ISL_678954\|20 | ···················· | ···················· | ···········T········· | ···················· | ···················· | ···················· |
| U | 2464 | hCoV-19/USA/MI-UM-10036686924/2020\|EPI_ISL_682101\| | ···················· | ···················· | ····················· | ···················· | ···················· | ···················· |
| U | 2465 | hCoV-19/USA/MI-UM-10036650310/2020\|EPI_ISL_682102\| | ···················· | ···················· | ····················· | ···················· | ···················· | ···················· |
| U | 2466 | hCoV-19/USA/MI-UM-10036710374/2020\|EPI_ISL_682103\| | ···················· | ···················· | ····················· | ···················· | ···················· | ···················· |
| U | 2467 | hCoV-19/USA/MI-UM-10036702303/2020\|EPI_ISL_682104\| | ···················· | ·A·················· | ····················· | ···················· | ···················· | ···················· |
| U | 2468 | hCoV-19/USA/MI-UM-10036740188/2020\|EPI_ISL_682105\| | ···················· | ···················· | ····················· | ···················· | ···················· | ···················· |
| U | 2469 | hCoV-19/USA/MI-UM-10036705555/2020\|EPI_ISL_682106\| | ···················· | ···················· | ····················· | ···················· | ···················· | ···················· |
| U | 2470 | hCoV-19/USA/MI-UM-10036706931/2020\|EPI_ISL_682107\| | ···················· | ···················· | ····················· | ···················· | ···················· | ···················· |
| U | 2471 | hCoV-19/USA/MI-UM-10036743706/2020\|EPI_ISL_682108\| | ···················· | ···················· | ····················· | ···················· | ···················· | ···················· |
| U | 2472 | hCoV-19/USA/MI-UM-10036706109/2020\|EPI_ISL_682109\| | ···················· | ·A·················· | ····················· | ···················· | ···················· | ···················· |
| U | 2473 | hCoV-19/USA/MI-UM-10036701359/2020\|EPI_ISL_682110\| | ···················· | ···················· | ····················· | ···················· | ···················· | ···················· |
| U | 2474 | hCoV-19/USA/MI-UM-10036742126/2020\|EPI_ISL_682111\| | ···················· | ···················· | ····················· | ···················· | ···················· | ···················· |
| U | 2475 | hCoV-19/USA/MI-UM-10036687887/2020\|EPI_ISL_682112\| | ···················· | ···················· | ····················· | ···················· | ···················· | ···················· |
| U | 2476 | hCoV-19/USA/MI-UM-10036707756/2020\|EPI_ISL_682113\| | ···················· | ·············A······ | ····················· | ···················· | ···················· | ···················· |
| U | 2477 | hCoV-19/USA/MI-UM-10036690451/2020\|EPI_ISL_682114\| | ···················· | ···················· | ····················· | ···················· | ···················· | ···················· |
| U | 2478 | hCoV-19/USA/MI-UM-10036686863/2020\|EPI_ISL_682115\| | ···················· | ···················· | ····················· | ···················· | ···················· | ···················· |
| U | 2479 | hCoV-19/USA/MI-UM-10036715361/2020\|EPI_ISL_682117\| | ···················· | ·A·················· | ····················· | ···················· | ···················· | ···················· |
| U | 2481 | hCoV-19/USA/MI-UM-10036689863/2020\|EPI_ISL_682119\| | ···················· | ···················· | ····················· | ···················· | ···················· | ···················· |
| U | 2484 | hCoV-19/USA/MI-UM-10036719444/2020\|EPI_ISL_682139\| | ···················· | ···················· | ····················· | ···················· | ···················· | ···················· |
| U | 2485 | hCoV-19/USA/MI-UM-10036711757/2020\|EPI_ISL_682140\| | ···················· | ···················· | ····················· | ···················· | ···················· | ···················· |
| U | 2486 | hCoV-19/USA/MI-UM-10036719494/2020\|EPI_ISL_682141\| | ···················· | ·A·················· | ····················· | ···················· | ···················· | ···················· |
| U | 2487 | hCoV-19/USA/MI-UM-10036704187/2020\|EPI_ISL_682143\| | ···················· | ·A·················· | ······G·············· | ···················· | ···················· | ···················· |

| | | | | | | | |
|---|---|---|---|---|---|---|---|
| U | 2488 hCoV-19/USA/MI-UM-10036721047/2020\|EPI_ISL_682144\| | ···················· | ·A·················· | ····················· | ···················· | ···················· | ···················· |
| U | 2489 hCoV-19/USA/MI-UM-10036707634/2020\|EPI_ISL_682146\| | ···················· | ···················· | ····················· | ···················· | ···················· | ···················· |
| U | 2490 hCoV-19/USA/MI-UM-10036708302/2020\|EPI_ISL_682147\| | ···················· | ···················· | ····················· | ···················· | ···················· | ···················· |
| U | 2491 hCoV-19/USA/MI-UM-10036710941/2020\|EPI_ISL_682148\| | ···················· | ···················· | ····················· | ···················· | ···················· | ···················· |
| U | 2492 hCoV-19/USA/MI-UM-10036720582/2020\|EPI_ISL_682149\| | ···················· | ·············A······ | ····················· | ···················· | ···················· | ···················· |
| U | 2497 hCoV-19/USA/MI-UM-10036721126/2020\|EPI_ISL_682158\| | ···················· | ···················· | ····················· | ···················· | ···················· | ···················· |
| U | 2498 hCoV-19/USA/MI-UM-10036721521/2020\|EPI_ISL_682159\| | ···················· | ···················· | ····················· | ···················· | ···················· | ···················· |
| U | 2499 hCoV-19/USA/MI-UM-10036721543/2020\|EPI_ISL_682160\| | ···················· | ······A············· | ····················· | ···················· | ···················· | ···················· |
| U | 2500 hCoV-19/USA/MI-UM-10036738304/2020\|EPI_ISL_682161\| | ···················· | ···················· | ····················· | ···················· | ···················· | ···················· |
| U | 2501 hCoV-19/USA/MI-UM-10036736854/2020\|EPI_ISL_682162\| | ···················· | ···················· | ····················· | ···················· | ···················· | ···················· |
| U | 2502 hCoV-19/USA/MI-UM-10036737267/2020\|EPI_ISL_682163\| | ···················· | ·············A······ | ····················· | ···················· | ···················· | ···················· |
| U | 2503 hCoV-19/USA/MI-UM-10036737474/2020\|EPI_ISL_682164\| | ···················· | ···················· | ····················· | ···················· | ···················· | ···················· |
| U | 2504 hCoV-19/USA/MI-UM-10036741616/2020\|EPI_ISL_682165\| | ···················· | ···················· | ····················· | ···················· | ···················· | ···················· |
| U | 2505 hCoV-19/USA/MI-UM-10036742180/2020\|EPI_ISL_682166\| | ···················· | ···················· | ····················· | ···················· | ···················· | ···················· |
| U | 2506 hCoV-19/USA/MI-UM-10036742402/2020\|EPI_ISL_682167\| | ···················· | ···················· | ····················· | ···················· | ···················· | ···················· |
| U | 2509 hCoV-19/USA/MI-UM-10036746274/2020\|EPI_ISL_682170\| | ···················· | ·A·················· | ····················· | ···················· | ···················· | ···A················ |
| U | 2510 hCoV-19/USA/MI-UM-10036744166/2020\|EPI_ISL_682171\| | ···················· | ···················· | ····················· | ···················· | ···················· | ···················· |
| U | 2511 hCoV-19/USA/MI-UM-10036760951/2020\|EPI_ISL_682172\| | ···················· | ······A············· | ····················· | ···················· | ···················· | ···················· |
| U | 2512 hCoV-19/USA/MI-UM-10036761214/2020\|EPI_ISL_682173\| | ···················· | ······A············· | ····················· | ···················· | ···················· | ···················· |
| U | 2513 hCoV-19/USA/MI-UM-10036743283/2020\|EPI_ISL_682174\| | ···················· | ···················· | ····················· | ···················· | ···················· | ···················· |
| U | 2514 hCoV-19/USA/MI-UM-10036756404/2020\|EPI_ISL_682175\| | ···················· | ·············A······ | ····················· | ···················· | ···················· | ···················· |
| U | 2515 hCoV-19/USA/MI-UM-10036745763/2020\|EPI_ISL_682177\| | ···················· | ···················· | ····················· | ···················· | ···················· | ···················· |
| U | 2516 hCoV-19/USA/MI-UM-10036743097/2020\|EPI_ISL_682178\| | ···················· | ···················· | ····················· | ···················· | ···················· | ···················· |
| U | 2517 hCoV-19/USA/MI-UM-10036750501/2020\|EPI_ISL_682179\| | ···················· | ···················· | ····················· | ···················· | ···················· | ···················· |
| U | 2518 hCoV-19/USA/MI-UM-10036761147/2020\|EPI_ISL_682180\| | ···················· | ···················· | ····················· | ···················· | ···················· | ···················· |
| U | 2519 hCoV-19/USA/MI-UM-10036761246/2020\|EPI_ISL_682181\| | ···················· | ···················· | ····················· | ···················· | ···················· | ···················· |
| U | 2520 hCoV-19/USA/MI-UM-10036741863/2020\|EPI_ISL_682182\| | ···················· | ···················· | ····················· | ···················· | ···················· | ···················· |
| U | 2521 hCoV-19/USA/MI-UM-10036744753/2020\|EPI_ISL_682183\| | ···················· | ···················· | ····················· | ···················· | ···················· | ···················· |
| U | 2522 hCoV-19/USA/MI-UM-10036746963/2020\|EPI_ISL_682184\| | ···················· | ···················· | ····················· | ···················· | ···················· | ···················· |
| U | 2523 hCoV-19/USA/MI-UM-10036757137/2020\|EPI_ISL_682185\| | ···················· | ···················· | ····················· | ···················· | ···················· | ···················· |
| U | 2524 hCoV-19/USA/MI-UM-10036747057/2020\|EPI_ISL_682187\| | ···················· | ·A·················· | ····················· | ···················· | ···················· | ···················· |
| U | 2525 hCoV-19/USA/MI-UM-10036738261/2020\|EPI_ISL_682188\| | ···················· | ···················· | ····················· | ···················· | ···················· | ···················· |
| U | 2526 hCoV-19/USA/MI-UM-10036757814/2020\|EPI_ISL_682189\| | ···················· | ···················· | ····················· | ···················· | ···················· | ···················· |
| U | 2527 hCoV-19/USA/MI-UM-10036761281/2020\|EPI_ISL_682190\| | ···················· | ···················· | ····················· | ···················· | ···················· | ···················· |
| U | 2528 hCoV-19/USA/MI-UM-10036743071/2020\|EPI_ISL_682191\| | ···················· | ···················· | ····················· | ···················· | ···················· | ···················· |
| U | 2529 hCoV-19/USA/MI-UM-10036746277/2020\|EPI_ISL_682192\| | ···················· | ·A·················· | ····················· | ···················· | ···················· | ···················· |
| U | 2531 hCoV-19/USA/MI-UM-10036743738/2020\|EPI_ISL_682194\| | ···················· | ···················· | ····················· | ···················· | ···················· | ···················· |
| U | 2532 hCoV-19/USA/MI-UM-10036746113/2020\|EPI_ISL_682195\| | ···················· | ···················· | ····················· | ···················· | ···················· | ···················· |
| U | 2533 hCoV-19/USA/MI-UM-10036765284/2020\|EPI_ISL_682196\| | ···················· | ···················· | ····················· | ···················· | ···················· | ···················· |
| U | 2534 hCoV-19/USA/MI-UM-10036767208/2020\|EPI_ISL_682197\| | ···················· | ···················· | ····················· | ···················· | ···················· | ···················· |
| U | 2537 hCoV-19/France/ARA-98691/2020\|EPI_ISL_683400\|2020- | ···················· | ···················· | ····················· | ···················· | ···················· | ···················· |
| U | 2538 hCoV-19/France/ARA-98746/2020\|EPI_ISL_683401\|2020- | ···················· | ···················· | ····················· | ···················· | ···················· | ···················· |
| U | 2540 hCoV-19/USA/TX-DSHS-1528/2020\|EPI_ISL_683443\|2020- | ···················· | ···················· | ····················· | ···················· | ···················· | ···················· |
| U | 2541 hCoV-19/Armenia/ChVir8659/2020\|EPI_ISL_683449\|2020 | ···················· | ···················· | ····················· | ···················· | ···················· | ···················· |
| U | 2542 hCoV-19/England/204780321/2020\|EPI_ISL_683478\|2020 | ···················· | ···················· | ····················· | ···················· | ···················· | ···················· |
| U | 2543 hCoV-19/England/204640953/2020\|EPI_ISL_683481\|2020 | ···················· | ···················· | ····················· | ···················· | ···················· | ···················· |
| U | 2544 hCoV-19/England/204790592/2020\|EPI_ISL_683482\|2020 | ···················· | ···················· | ····················· | ···················· | ···················· | ···················· |
| U | 2545 hCoV-19/England/204820616/2020\|EPI_ISL_683483\|2020 | ···················· | ···················· | ····················· | ···················· | ···················· | ···················· |
| U | 2546 hCoV-19/England/C200046845/2020\|EPI_ISL_683485\|202 | ···················· | ···················· | ····················· | ···················· | ···················· | ···················· |
| U | 2547 hCoV-19/England/C200046933/2020\|EPI_ISL_683486\|202 | ···················· | ···················· | ····················· | ···················· | ···················· | ···················· |
| U | 2548 hCoV-19/England/C200046950/2020\|EPI_ISL_683487\|202 | ···················· | ···················· | ····················· | ···················· | ···················· | ···················· |
| U | 2549 hCoV-19/England/C200046965/2020\|EPI_ISL_683488\|202 | ···················· | ···················· | ····················· | ···················· | ···················· | ···················· |
| U | 2550 hCoV-19/England/C200049134/2020\|EPI_ISL_683489\|202 | ···················· | ···················· | ····················· | ···················· | ···················· | ···················· |
| U | 2551 hCoV-19/England/C200049138/2020\|EPI_ISL_683490\|202 | ···················· | ···················· | ····················· | ···················· | ···················· | ···················· |
| U | 2552 hCoV-19/England/204561377/2020\|EPI_ISL_683491\|2020 | ···················· | ···················· | ····················· | ···················· | ···················· | ···················· |
| U | 2553 hCoV-19/England/204582194/2020\|EPI_ISL_683492\|2020 | ···················· | ···················· | ····················· | ···················· | ···················· | ···················· |
| U | 2554 hCoV-19/England/204590427/2020\|EPI_ISL_683493\|2020 | ···················· | ···················· | ····················· | ···················· | ···················· | ···················· |
| U | 2555 hCoV-19/England/204590444/2020\|EPI_ISL_683494\|2020 | ···················· | ···················· | ····················· | ···················· | ···················· | ···················· |
| U | 2556 hCoV-19/England/204590445/2020\|EPI_ISL_683495\|2020 | ···················· | ···················· | ····················· | ···················· | ···················· | ···················· |
| U | 2557 hCoV-19/England/204590447/2020\|EPI_ISL_683496\|2020 | ···················· | ···················· | ····················· | ···················· | ···················· | ···················· |
| U | 2558 hCoV-19/England/204590450/2020\|EPI_ISL_683497\|2020 | ···················· | ···················· | ····················· | ···················· | ···················· | ···················· |
| U | 2559 hCoV-19/England/204590451/2020\|EPI_ISL_683498\|2020 | ···················· | ···················· | ····················· | ···················· | ···················· | ···················· |
| U | 2560 hCoV-19/England/204590452/2020\|EPI_ISL_683499\|2020 | ···················· | ···················· | ····················· | ···················· | ···················· | ···················· |
| U | 2561 hCoV-19/Australia/WA407-2/2020\|EPI_ISL_672619\|2020 | ···················· | ···················· | ····················· | ···················· | ···················· | ···················· |
| U | 2562 hCoV-19/Australia/WA409/2020\|EPI_ISL_672621\|2020-0 | ···················· | ···················· | ····················· | ···················· | ···················· | ···················· |
| U | 2563 hCoV-19/Australia/WA411/2020\|EPI_ISL_672623\|2020-0 | ···················· | ···················· | ····················· | ···················· | ···················· | ···················· |
| U | 2564 hCoV-19/Australia/WA413/2020\|EPI_ISL_672625\|2020-0 | ···················· | ···················· | ····················· | ···················· | ···················· | ···················· |
| U | 2565 hCoV-19/Australia/WA414/2020\|EPI_ISL_672626\|2020-0 | ···················· | ···················· | ····················· | ···················· | ···················· | ···················· |
| U | 2566 hCoV-19/Australia/WA418/2020\|EPI_ISL_672630\|2020-0 | ···················· | ···················· | ····················· | ···················· | ···················· | ···················· |
| U | 2567 hCoV-19/Australia/WA419/2020\|EPI_ISL_672631\|2020-0 | ···················· | ···················· | ····················· | ···················· | ···················· | ···················· |
| U | 2568 hCoV-19/Australia/WA421/2020\|EPI_ISL_672633\|2020-0 | ···················· | ···················· | ····················· | ···················· | ···················· | ···················· |
| U | 2569 hCoV-19/Australia/WA422/2020\|EPI_ISL_672634\|2020-0 | ···················· | ···················· | ····················· | ···················· | ···················· | ···················· |
| U | 2570 hCoV-19/Australia/WA423/2020\|EPI_ISL_672635\|2020-0 | ···················· | ···················· | ····················· | ···················· | ···················· | ···················· |
| U | 2571 hCoV-19/Australia/WA425/2020\|EPI_ISL_672637\|2020-0 | ···················· | ···················· | ····················· | ···················· | ···················· | ···················· |
| U | 2572 hCoV-19/Australia/WA430/2020\|EPI_ISL_672642\|2020-0 | ···················· | ···················· | ····················· | ···················· | ···················· | ···················· |
| U | 2573 hCoV-19/Australia/WA432/2020\|EPI_ISL_672644\|2020-0 | ···················· | ···················· | ····················· | ···················· | ···················· | ···················· |
| U | 2574 hCoV-19/Australia/WA435/2020\|EPI_ISL_672647\|2020-0 | ···················· | ···················· | ····················· | ···················· | ···················· | ···················· |
| U | 2575 hCoV-19/Australia/WA436/2020\|EPI_ISL_672648\|2020-0 | ···················· | ···················· | ····················· | ···················· | ···················· | ···················· |
| U | 2576 hCoV-19/Australia/WA438/2020\|EPI_ISL_672650\|2020-0 | ···················· | ···················· | ····················· | ···················· | ···················· | ···················· |
| U | 2577 hCoV-19/Australia/WA440/2020\|EPI_ISL_672652\|2020-0 | ···················· | ···················· | ····················· | ···················· | ···················· | ···················· |

| | | | | | | | | |
|---|---|---|---|---|---|---|---|---|
| U | 2578 | hCoV-19/Australia/WA441/2020\|EPI_ISL_672653\|2020-0 | ···················· | ·A·················· | ····················· | ···················· | ···················· | ···A················ |
| U | 2579 | hCoV-19/Australia/NSW1211/2020\|EPI_ISL_672655\|2020 | ···················· | ···················· | ····················· | ···················· | ···················· | ···················· |
| U | 2580 | hCoV-19/Australia/NSW1212/2020\|EPI_ISL_672656\|2020 | ···················· | ·A·················· | ····················· | ···················· | ···················· | ···················· |
| U | 2581 | hCoV-19/Australia/NSW1213/2020\|EPI_ISL_672657\|2020 | ···················· | ·A·················· | ····················· | ···················· | ···················· | ···················· |
| U | 2582 | hCoV-19/Australia/NSW1214/2020\|EPI_ISL_672658\|2020 | ···················· | ···················· | ····················· | ···················· | ···················· | ···················· |
| U | 2583 | hCoV-19/Australia/NSW1215/2020\|EPI_ISL_672659\|2020 | ···················· | ·A·················· | ····················· | ···················· | ···················· | ···A················ |
| U | 2584 | hCoV-19/Australia/NSW1216/2020\|EPI_ISL_672660\|2020 | ···················· | ·A·················· | ····················· | ···················· | ···················· | ···A················ |
| U | 2585 | hCoV-19/Australia/NSW1217/2020\|EPI_ISL_672661\|2020 | ···················· | ···················· | ····················· | ···················· | ···················· | ···················· |
| U | 2586 | hCoV-19/Australia/NSW1218/2020\|EPI_ISL_672662\|2020 | ···················· | ···················· | ····················· | ···················· | ···················· | ···················· |
| U | 2587 | hCoV-19/Brazil/SP-L25-CD635/2020\|EPI_ISL_672705\|20 | ···················· | ···················· | ····················· | ···················· | ···················· | ···················· |
| U | 2588 | hCoV-19/Brazil/SP-L25-CD647/2020\|EPI_ISL_672711\|20 | ···················· | ···················· | ····················· | ···················· | ···················· | ···················· |
| U | 2589 | hCoV-19/Brazil/SP-L25-CD669/2020\|EPI_ISL_672719\|20 | ···················· | ···················· | ····················· | ···················· | ···················· | ···················· |
| U | 2590 | hCoV-19/Brazil/SP-L25-CD670/2020\|EPI_ISL_672720\|20 | ···················· | ···················· | ····················· | ···················· | ···················· | ···················· |
| U | 2591 | hCoV-19/England/MILK-B8676C/2020\|EPI_ISL_673941\|20 | ···················· | ···················· | ····················· | ···················· | ···················· | ···················· |
| U | 2592 | hCoV-19/England/MILK-B86034/2020\|EPI_ISL_673943\|20 | ···················· | ···················· | ····················· | ···················· | ···················· | ···················· |
| U | 2593 | hCoV-19/England/MILK-B8640B/2020\|EPI_ISL_673944\|20 | ···················· | ···················· | ····················· | ···················· | ···················· | ···················· |
| U | 2594 | hCoV-19/England/MILK-B8D56B/2020\|EPI_ISL_673945\|20 | ···················· | ···················· | ····················· | ···················· | ···················· | ···················· |
| U | 2595 | hCoV-19/England/MILK-B8D5D4/2020\|EPI_ISL_673946\|20 | ···················· | ···················· | ····················· | ···················· | ···················· | ···················· |
| U | 2596 | hCoV-19/England/MILK-B8D659/2020\|EPI_ISL_673949\|20 | ···················· | ···················· | ····················· | ···················· | ···················· | ···················· |
| U | 2597 | hCoV-19/England/MILK-B85E3B/2020\|EPI_ISL_673951\|20 | ···················· | ···················· | ····················· | ···················· | ···················· | ···················· |
| U | 2598 | hCoV-19/England/MILK-B8E7FB/2020\|EPI_ISL_673952\|20 | ···················· | ···················· | ····················· | ···················· | ···················· | ···················· |
| U | 2599 | hCoV-19/England/MILK-B8D2FB/2020\|EPI_ISL_673953\|20 | ···················· | ···················· | ····················· | ···················· | ···················· | ···················· |
| U | 2600 | hCoV-19/England/MILK-B8691B/2020\|EPI_ISL_673954\|20 | ···················· | ···················· | ····················· | ···················· | ···················· | ···················· |
| U | 2601 | hCoV-19/England/MILK-B8619B/2020\|EPI_ISL_673955\|20 | ···················· | ···················· | ····················· | ···················· | ···················· | ···················· |
| U | 2602 | hCoV-19/England/MILK-B860E9/2020\|EPI_ISL_673957\|20 | ···················· | ···················· | ····················· | ···················· | ···················· | ···················· |
| U | 2603 | hCoV-19/England/MILK-B863E0/2020\|EPI_ISL_673958\|20 | ···················· | ···················· | ····················· | ···················· | ···················· | ···················· |
| U | 2604 | hCoV-19/England/MILK-B8692A/2020\|EPI_ISL_673959\|20 | ···················· | ···················· | ····················· | ···················· | ···················· | ···················· |
| U | 2605 | hCoV-19/England/MILK-B8616E/2020\|EPI_ISL_673961\|20 | ···················· | ···················· | ····················· | ···················· | ···················· | ···················· |
| U | 2606 | hCoV-19/England/MILK-B85BCB/2020\|EPI_ISL_673963\|20 | ···················· | ···················· | ····················· | ···················· | ···················· | ···················· |
| U | 2607 | hCoV-19/England/MILK-B8E6B2/2020\|EPI_ISL_673964\|20 | ···················· | ···················· | ····················· | ···················· | ···················· | ···················· |
| U | 2608 | hCoV-19/England/MILK-B86289/2020\|EPI_ISL_673965\|20 | ···················· | ···················· | ····················· | ···················· | ···················· | ···················· |
| U | 2609 | hCoV-19/England/MILK-B8E597/2020\|EPI_ISL_673966\|20 | ···················· | ···················· | ····················· | ···················· | ···················· | ···················· |
| U | 2610 | hCoV-19/England/MILK-B8683C/2020\|EPI_ISL_673967\|20 | ···················· | ·A·················· | ····················· | ···················· | ···················· | ···A················ |
| U | 2611 | hCoV-19/England/MILK-B85C22/2020\|EPI_ISL_673968\|20 | ···················· | ···················· | ····················· | ···················· | ···················· | ···················· |
| U | 2612 | hCoV-19/England/MILK-B8622F/2020\|EPI_ISL_673969\|20 | ···················· | ···················· | ····················· | ···················· | ···················· | ···················· |
| U | 2613 | hCoV-19/England/MILK-B85DE3/2020\|EPI_ISL_673970\|20 | ···················· | ···················· | ····················· | ···················· | ···················· | ···················· |
| U | 2614 | hCoV-19/England/MILK-B8E55B/2020\|EPI_ISL_673972\|20 | ···················· | ···················· | ····················· | ···················· | ···················· | ···················· |
| U | 2615 | hCoV-19/England/MILK-B8E70A/2020\|EPI_ISL_673973\|20 | ···················· | ···················· | ····················· | ···················· | ···················· | ···················· |
| U | 2616 | hCoV-19/England/MILK-B8609E/2020\|EPI_ISL_673974\|20 | ···················· | ···················· | ····················· | ···················· | ···················· | ···················· |
| U | 2617 | hCoV-19/England/MILK-B8632C/2020\|EPI_ISL_673975\|20 | ···················· | ···················· | ····················· | ···················· | ···················· | ···················· |
| U | 2618 | hCoV-19/England/MILK-B86007/2020\|EPI_ISL_673976\|20 | ···················· | ···················· | ····················· | ···················· | ···················· | ···················· |
| U | 2619 | hCoV-19/England/MILK-B8D5E3/2020\|EPI_ISL_673977\|20 | ···················· | ···················· | ····················· | ···················· | ···················· | ···················· |
| U | 2620 | hCoV-19/England/MILK-B85E0E/2020\|EPI_ISL_673978\|20 | ···················· | ···················· | ····················· | ···················· | ···················· | ···················· |
| U | 2621 | hCoV-19/England/MILK-B8D4C8/2020\|EPI_ISL_673979\|20 | ···················· | ···················· | ····················· | ···················· | ···················· | ···················· |
| U | 2622 | hCoV-19/England/MILK-B8E5B5/2020\|EPI_ISL_673981\|20 | ···················· | ·A·················· | ····················· | ···················· | ···················· | ···A················ |
| U | 2623 | hCoV-19/England/MILK-B8D48C/2020\|EPI_ISL_673982\|20 | ···················· | ···················· | ····················· | ···················· | ···················· | ···················· |
| U | 2624 | hCoV-19/England/MILK-B85F47/2020\|EPI_ISL_673983\|20 | ···················· | ···················· | ····················· | ···················· | ···················· | ···················· |
| U | 2625 | hCoV-19/England/MILK-B8673F/2020\|EPI_ISL_673984\|20 | ···················· | ···················· | ····················· | ···················· | ···················· | ···················· |
| U | 2626 | hCoV-19/England/MILK-B85F83/2020\|EPI_ISL_673985\|20 | ···················· | ···················· | ····················· | ···················· | ···················· | ···················· |
| U | 2627 | hCoV-19/England/MILK-B8D677/2020\|EPI_ISL_673986\|20 | ···················· | ···················· | ····················· | ···················· | ···················· | ···················· |
| U | 2628 | hCoV-19/England/MILK-B8D62C/2020\|EPI_ISL_673988\|20 | ···················· | ···················· | ····················· | ···················· | ···················· | ···················· |
| U | 2629 | hCoV-19/England/MILK-B863B3/2020\|EPI_ISL_673989\|20 | ···················· | ···················· | ····················· | ···················· | ···················· | ···················· |
| U | 2630 | hCoV-19/England/MILK-B8E7A0/2020\|EPI_ISL_673991\|20 | ···················· | ···················· | ····················· | ···················· | ···················· | ···················· |
| U | 2631 | hCoV-19/England/MILK-B8631D/2020\|EPI_ISL_673993\|20 | ···················· | ···················· | ····················· | ···················· | ···················· | ···················· |
| U | 2632 | hCoV-19/England/MILK-B8E6D0/2020\|EPI_ISL_673994\|20 | ···················· | ···················· | ····················· | ···················· | ···················· | ···················· |
| U | 2633 | hCoV-19/England/MILK-B8D3BC/2020\|EPI_ISL_673995\|20 | ···················· | ···················· | ····················· | ···················· | ···················· | ···················· |
| U | 2634 | hCoV-19/England/MILK-B864DE/2020\|EPI_ISL_673996\|20 | ···················· | ···················· | ····················· | ···················· | ···················· | ···················· |
| U | 2635 | hCoV-19/England/MILK-B8E6C1/2020\|EPI_ISL_673997\|20 | ···················· | ···················· | ····················· | ···················· | ···················· | ···················· |
| U | 2636 | hCoV-19/England/MILK-B96866/2020\|EPI_ISL_676200\|20 | ···················· | ···················· | ····················· | ···················· | ···················· | ···················· |
| U | 2637 | hCoV-19/England/MILK-B9424C/2020\|EPI_ISL_676201\|20 | ···················· | ···················· | ····················· | ···················· | ···················· | ···················· |
| U | 2638 | hCoV-19/England/MILK-B96101/2020\|EPI_ISL_676203\|20 | ···················· | ·A·················· | ····················· | ···················· | ···················· | ···A················ |
| U | 2639 | hCoV-19/England/MILK-B960AA/2020\|EPI_ISL_676204\|20 | ···················· | ···················· | ····················· | ···················· | ···················· | ···················· |
| U | 2640 | hCoV-19/England/MILK-B93D3D/2020\|EPI_ISL_676206\|20 | ···················· | ···················· | ····················· | ···················· | ···················· | ···················· |
| U | 2641 | hCoV-19/England/MILK-B94F36/2020\|EPI_ISL_676207\|20 | ···················· | ···················· | ····················· | ···················· | ···················· | ···················· |
| U | 2642 | hCoV-19/England/MILK-B93DC4/2020\|EPI_ISL_676208\|20 | ···················· | ···················· | ····················· | ···················· | ···················· | ···················· |
| U | 2643 | hCoV-19/England/MILK-B944BF/2020\|EPI_ISL_676209\|20 | ···················· | ···················· | ····················· | ···················· | ···················· | ···················· |
| U | 2644 | hCoV-19/England/MILK-B94446/2020\|EPI_ISL_676212\|20 | ···················· | ···················· | ····················· | ···················· | ···················· | ···················· |
| U | 2645 | hCoV-19/England/MILK-B93FFB/2020\|EPI_ISL_676213\|20 | ···················· | ···················· | ····················· | ···················· | ···················· | ···················· |
| U | 2646 | hCoV-19/England/MILK-B96ABB/2020\|EPI_ISL_676214\|20 | ···················· | ···················· | ····················· | ···················· | ···················· | ···················· |
| U | 2647 | hCoV-19/England/MILK-B96936/2020\|EPI_ISL_676215\|20 | ···················· | ···················· | ····················· | ···················· | ···················· | ···················· |
| U | 2648 | hCoV-19/England/MILK-B96417/2020\|EPI_ISL_676260\|20 | ···················· | ···················· | ····················· | ···················· | ···················· | ···················· |
| U | 2649 | hCoV-19/England/MILK-B94983/2020\|EPI_ISL_676262\|20 | ···················· | ···················· | ····················· | ···················· | ···················· | ···················· |
| U | 2650 | hCoV-19/England/MILK-B94895/2020\|EPI_ISL_676266\|20 | ···················· | ···················· | ····················· | ···················· | ···················· | ···················· |
| U | 2651 | hCoV-19/England/MILK-B944A0/2020\|EPI_ISL_676267\|20 | ···················· | ···················· | ····················· | ···················· | ···················· | ···················· |
| U | 2652 | hCoV-19/England/MILK-B943C1/2020\|EPI_ISL_676268\|20 | ···················· | ···················· | ····················· | ···················· | ···················· | ···················· |
| U | 2653 | hCoV-19/England/MILK-B94EC0/2020\|EPI_ISL_676269\|20 | ···················· | ···················· | ····················· | ···················· | ···················· | ···················· |
| U | 2654 | hCoV-19/England/MILK-B9670F/2020\|EPI_ISL_676271\|20 | ···················· | ·A·················· | ····················· | ···················· | ···················· | ···A················ |
| U | 2655 | hCoV-19/England/MILK-B94482/2020\|EPI_ISL_676272\|20 | ···················· | ·R·················· | ····················· | ···················· | ···················· | ···R················ |
| U | 2656 | hCoV-19/England/MILK-B94F81/2020\|EPI_ISL_676273\|20 | ···················· | ···················· | ····················· | ···················· | ···················· | ···················· |
| U | 2657 | hCoV-19/England/MILK-B95D59/2020\|EPI_ISL_676274\|20 | ···················· | ···················· | ····················· | ···················· | ···················· | ···················· |

| | | | | | | | | |
|---|---|---|---|---|---|---|---|---|
| U | 2658 | hCoV-19/England/MILK-B942D3/2020\|EPI_ISL_676275\|20 | ···················· | ···················· | ···················· | ···················· | ···················· | ···················· |
| U | 2659 | hCoV-19/England/MILK-B94974/2020\|EPI_ISL_676276\|20 | ···················· | ···················· | ···················· | ···················· | ···················· | ···················· |
| U | 2660 | hCoV-19/England/MILK-B96383/2020\|EPI_ISL_676280\|20 | ···················· | ·A·················· | ···················· | ···················· | ···················· | ···A················ |
| U | 2661 | hCoV-19/England/MILK-B968EE/2020\|EPI_ISL_676281\|20 | ···················· | ···················· | ···················· | ···················· | ···················· | ···················· |
| U | 2662 | hCoV-19/England/MILK-B95FCC/2020\|EPI_ISL_676283\|20 | ···················· | ···················· | ···················· | ···················· | ···················· | ···················· |
| U | 2663 | hCoV-19/England/MILK-B9613E/2020\|EPI_ISL_676284\|20 | ···················· | ···················· | ···················· | ···················· | ···················· | ···················· |
| U | 2664 | hCoV-19/England/MILK-B939FD/2020\|EPI_ISL_676285\|20 | ···················· | ···················· | ···················· | ···················· | ···················· | ···················· |
| U | 2665 | hCoV-19/England/MILK-B96110/2020\|EPI_ISL_676286\|20 | ···················· | ·A·················· | ···················· | ···················· | ···················· | ···A················ |
| U | 2666 | hCoV-19/England/MILK-B969EB/2020\|EPI_ISL_676287\|20 | ···················· | ···················· | ···················· | ···················· | ···················· | ···················· |
| U | 2667 | hCoV-19/England/MILK-B94CC6/2020\|EPI_ISL_676288\|20 | ···················· | ·A·················· | ···················· | ···················· | ···················· | ···················· |
| U | 2668 | hCoV-19/England/MILK-B94288/2020\|EPI_ISL_676290\|20 | ···················· | ···················· | ···················· | ···················· | ···················· | ···················· |
| U | 2669 | hCoV-19/England/MILK-B95FDB/2020\|EPI_ISL_676291\|20 | ···················· | ···················· | ···················· | ···················· | ···················· | ···················· |
| U | 2670 | hCoV-19/England/MILK-B943B2/2020\|EPI_ISL_676292\|20 | ···················· | ···················· | ···················· | ···················· | ···················· | ···················· |
| U | 2671 | hCoV-19/England/MILK-B94D78/2020\|EPI_ISL_676293\|20 | ···················· | ···················· | ···················· | ···················· | ···················· | ···················· |
| U | 2672 | hCoV-19/England/MILK-B96408/2020\|EPI_ISL_676294\|20 | ···················· | ···················· | ···················· | ···················· | ···················· | ···················· |
| U | 2673 | hCoV-19/England/MILK-B9673C/2020\|EPI_ISL_676295\|20 | ···················· | ···················· | ···················· | ···················· | ···················· | ···················· |
| U | 2674 | hCoV-19/England/MILK-B9664E/2020\|EPI_ISL_676296\|20 | ···················· | ···················· | ···················· | ···················· | ···················· | ···················· |
| U | 2675 | hCoV-19/England/MILK-B96338/2020\|EPI_ISL_676297\|20 | ···················· | ···················· | ···················· | ···················· | ···················· | ···················· |
| U | 2676 | hCoV-19/England/MILK-B948B3/2020\|EPI_ISL_676298\|20 | ···················· | ···················· | ···················· | ···················· | ······T············· | ···················· |
| U | 2677 | hCoV-19/England/MILK-B93957/2020\|EPI_ISL_676299\|20 | ···················· | ···················· | ···················· | ···················· | ···················· | ···················· |
| U | 2678 | hCoV-19/USA/WI-UW-2095/2020\|EPI_ISL_677500\|2020-11 | ···················· | ···················· | ···················· | ···················· | ···················· | ···················· |
| U | 2684 | hCoV-19/USA/WI-UW-2101/2020\|EPI_ISL_677506\|2020-11 | ···················· | ···················· | ···················· | ···················· | ···················· | ···················· |
| U | 2686 | hCoV-19/USA/WI-UW-2103/2020\|EPI_ISL_677508\|2020-11 | ···················· | ···················· | ···················· | ···················· | ···················· | ···················· |
| U | 2696 | hCoV-19/USA/WI-UW-2114/2020\|EPI_ISL_677519\|2020-10 | ···················· | ···················· | ···················· | ···················· | ···················· | ···················· |
| U | 2698 | hCoV-19/USA/WI-UW-2117/2020\|EPI_ISL_677522\|2020-10 | ···················· | ···················· | ···················· | ···················· | ···················· | ···················· |
| U | 2703 | hCoV-19/USA/WI-UW-2122/2020\|EPI_ISL_677527\|2020-09 | ···················· | ···················· | ···················· | ···················· | ···················· | ···················· |
| U | 2710 | hCoV-19/USA/IL-UW-2130/2020\|EPI_ISL_677535\|2020-09 | ···················· | ···················· | ···················· | ···················· | ···················· | ···················· |
| U | 2716 | hCoV-19/USA/WI-UW-2188/2020\|EPI_ISL_677591\|2020-10 | ···················· | ···················· | ···················· | ···················· | ···················· | ···················· |
| U | 2718 | hCoV-19/USA/WI-UW-2190/2020\|EPI_ISL_677593\|2020-10 | ···················· | ···················· | ···················· | ···················· | ···················· | ···················· |
| U | 2720 | hCoV-19/USA/WI-UW-2192/2020\|EPI_ISL_677595\|2020-10 | ···················· | ···················· | ···················· | ···················· | ···················· | ···················· |
| U | 2721 | hCoV-19/USA/WI-UW-2193/2020\|EPI_ISL_677596\|2020-10 | ···················· | ···················· | ···················· | ···················· | ···················· | ···················· |
| U | 2722 | hCoV-19/USA/WI-UW-2194/2020\|EPI_ISL_677597\|2020-10 | ···················· | ···················· | ···················· | ···················· | ···················· | ···················· |
| U | 2723 | hCoV-19/USA/WI-UW-2195/2020\|EPI_ISL_677598\|2020-10 | ···················· | ···················· | ···················· | ···················· | ···················· | ···················· |
| U | 2725 | hCoV-19/England/204881275/2020\|EPI_ISL_678805\|2020 | ···················· | ···················· | ···················· | ···················· | ···················· | ···················· |
| U | 2726 | hCoV-19/England/204881291/2020\|EPI_ISL_678813\|2020 | ···················· | ···················· | ···················· | ···················· | ···················· | ···················· |
| U | 2727 | hCoV-19/England/204920938/2020\|EPI_ISL_678828\|2020 | ···················· | ···················· | ···················· | ···················· | ···················· | ···················· |
| U | 2728 | hCoV-19/England/ALDP-B5CD4F/2020\|EPI_ISL_678834\|20 | ···················· | ···················· | ···················· | ···················· | ···················· | ···A················ |
| U | 2729 | hCoV-19/England/ALDP-B5CD5E/2020\|EPI_ISL_678835\|20 | ···················· | ···················· | ···················· | ···················· | ···················· | ···················· |
| U | 2730 | hCoV-19/England/ALDP-B5CD6D/2020\|EPI_ISL_678836\|20 | ···················· | ···················· | ···················· | ···················· | ···················· | ···················· |
| U | 2731 | hCoV-19/England/ALDP-B5CD7C/2020\|EPI_ISL_678837\|20 | ···················· | ···················· | ···················· | ···················· | ···················· | ···················· |
| U | 2732 | hCoV-19/England/ALDP-B5CFC1/2020\|EPI_ISL_678840\|20 | ···················· | ···················· | ···················· | ···················· | ······T············· | ···················· |
| U | 2733 | hCoV-19/England/ALDP-B5CFD0/2020\|EPI_ISL_678841\|20 | ···················· | ···················· | ···················· | ···················· | ···················· | ···················· |
| U | 2734 | hCoV-19/England/ALDP-B5CFEF/2020\|EPI_ISL_678842\|20 | ···················· | ···················· | ···················· | ···················· | ···················· | ···················· |
| U | 2735 | hCoV-19/England/ALDP-B5CFFE/2020\|EPI_ISL_678843\|20 | ···················· | ···················· | ···················· | ···················· | ······T············· | ···················· |
| U | 2736 | hCoV-19/England/ALDP-B5D018/2020\|EPI_ISL_678844\|20 | ···················· | ···················· | ···················· | ···················· | ···················· | ···················· |
| U | 2737 | hCoV-19/England/ALDP-B5D027/2020\|EPI_ISL_678845\|20 | ···················· | ···················· | ···················· | ···················· | ···················· | ···················· |
| U | 2738 | hCoV-19/England/ALDP-B5D063/2020\|EPI_ISL_678849\|20 | ···················· | ···················· | ···················· | ···················· | ···················· | ···················· |
| U | 2739 | hCoV-19/England/ALDP-B5D0BE/2020\|EPI_ISL_678853\|20 | ···················· | ···················· | ···················· | ···················· | ···················· | ···················· |
| U | 2740 | hCoV-19/England/ALDP-B5D0CD/2020\|EPI_ISL_678854\|20 | ···················· | ···················· | ···················· | ···················· | ···················· | ···················· |
| U | 2741 | hCoV-19/England/ALDP-B5D106/2020\|EPI_ISL_678857\|20 | ···················· | ···················· | ···················· | ···················· | ···················· | ···················· |
| U | 2742 | hCoV-19/England/ALDP-B5D115/2020\|EPI_ISL_678858\|20 | ···················· | ···················· | ···················· | ···················· | ···················· | ···················· |
| U | 2743 | hCoV-19/England/ALDP-B5D124/2020\|EPI_ISL_678859\|20 | ···················· | ···················· | ···················· | ···················· | ···················· | ···················· |
| U | 2744 | hCoV-19/England/ALDP-B5D133/2020\|EPI_ISL_678860\|20 | ···················· | ···················· | ···················· | ···················· | ···················· | ···················· |
| U | 2745 | hCoV-19/England/ALDP-B5D151/2020\|EPI_ISL_678862\|20 | ···················· | ···················· | ···················· | ···················· | ···················· | ···················· |
| U | 2746 | hCoV-19/England/ALDP-B5D17F/2020\|EPI_ISL_678863\|20 | ···················· | ···················· | ···················· | ···················· | ···················· | ···················· |
| U | 2747 | hCoV-19/England/ALDP-B5D18E/2020\|EPI_ISL_678864\|20 | ···················· | ···················· | ···················· | ···················· | ···················· | ···················· |
| U | 2748 | hCoV-19/England/ALDP-B5D19D/2020\|EPI_ISL_678865\|20 | ···················· | ···················· | ···················· | ···················· | ···················· | ···················· |
| U | 2749 | hCoV-19/England/ALDP-B5D1BB/2020\|EPI_ISL_678867\|20 | ···················· | ·A·················· | ···················· | ···················· | ···················· | ···A················ |
| U | 2750 | hCoV-19/England/ALDP-B5D1D9/2020\|EPI_ISL_678869\|20 | ···················· | ···················· | ···················· | ···················· | ···················· | ···················· |
| U | 2751 | hCoV-19/England/ALDP-B5D24F/2020\|EPI_ISL_678876\|20 | ···················· | ···················· | ···················· | ···················· | ···················· | ···················· |
| U | 2752 | hCoV-19/England/ALDP-B5D25E/2020\|EPI_ISL_678877\|20 | ···················· | ···················· | ···················· | ···················· | ···················· | ···················· |
| U | 2753 | hCoV-19/USA/PA-MGEL-00326/2020\|EPI_ISL_682000\|2020 | ···················· | ···················· | ···················· | ···················· | ···················· | ···················· |
| U | 2754 | hCoV-19/USA/PA-MGEL-00327/2020\|EPI_ISL_682001\|2020 | ···················· | ···················· | ···················· | ···················· | ···················· | ···················· |
| U | 2755 | hCoV-19/USA/PA-MGEL-00328/2020\|EPI_ISL_682002\|2020 | ···················· | ···················· | ···················· | ···················· | ···················· | ···················· |
| U | 2756 | hCoV-19/USA/PA-MGEL-00329/2020\|EPI_ISL_682003\|2020 | ···················· | ···················· | ···················· | ···················· | ···················· | ···················· |
| U | 2757 | hCoV-19/USA/PA-MGEL-00331/2020\|EPI_ISL_682004\|2020 | ···················· | ···················· | ···················· | ···················· | ···················· | ···················· |
| U | 2758 | hCoV-19/USA/PA-MGEL-00396/2020\|EPI_ISL_682005\|2020 | ···················· | ···················· | ···················· | ···················· | ···················· | ···················· |
| U | 2759 | hCoV-19/USA/PA-MGEL-00397/2020\|EPI_ISL_682006\|2020 | ···················· | ···················· | ···················· | ···················· | ···················· | ···················· |
| U | 2760 | hCoV-19/USA/PA-MGEL-00398/2020\|EPI_ISL_682007\|2020 | ···················· | ···················· | ···················· | ···················· | ···················· | ···················· |
| U | 2761 | hCoV-19/USA/PA-MGEL-00400/2020\|EPI_ISL_682008\|2020 | ···················· | ···················· | ···················· | ···················· | ···················· | ···················· |
| U | 2762 | hCoV-19/USA/PA-MGEL-00401/2020\|EPI_ISL_682009\|2020 | ···················· | ···················· | ···················· | ···················· | ···················· | ···················· |
| U | 2763 | hCoV-19/USA/PA-MGEL-00403/2020\|EPI_ISL_682010\|2020 | ···················· | ···················· | ···················· | ···················· | ···················· | ···················· |
| U | 2764 | hCoV-19/USA/PA-MGEL-00404/2020\|EPI_ISL_682011\|2020 | ···················· | ···················· | ···················· | ···················· | ···················· | ···················· |
| U | 2765 | hCoV-19/USA/PA-MGEL-00406/2020\|EPI_ISL_682012\|2020 | ···················· | ···················· | ···················· | ···················· | ···················· | ···················· |
| U | 2766 | hCoV-19/USA/MD-MDH-0457/2020\|EPI_ISL_682068\|2020-1 | ···················· | ···················· | ···················· | ···················· | ···················· | ···················· |
| U | 2767 | hCoV-19/USA/MD-MDH-0458/2020\|EPI_ISL_682069\|2020-1 | ···················· | ···················· | ···················· | ···················· | ···················· | ···················· |
| U | 2768 | hCoV-19/USA/MD-MDH-0459/2020\|EPI_ISL_682070\|2020-1 | ···················· | ···················· | ···················· | ···················· | ···················· | ···················· |
| U | 2769 | hCoV-19/USA/MD-MDH-0460/2020\|EPI_ISL_682071\|2020-1 | ···················· | ············C······· | ···················· | ···················· | ···················· | ···················· |
| U | 2770 | hCoV-19/USA/MD-MDH-0461/2020\|EPI_ISL_682072\|2020-1 | ···················· | ···················· | ···················· | ···················· | ···················· | ···················· |
| U | 2771 | hCoV-19/USA/MD-MDH-0462/2020\|EPI_ISL_682073\|2020-1 | ···················· | ···················· | ···················· | ···················· | ···················· | ···················· |

| | | | | | | | | |
|---|---|---|---|---|---|---|---|---|
| U | 2772 | hCoV-19/USA/MD-MDH-0463/2020\|EPI_ISL_682074\|2020-1 | ···················· | ···················· | ···················· | ···················· | ···················· | ···················· |
| U | 2773 | hCoV-19/USA/MD-MDH-0464/2020\|EPI_ISL_682075\|2020-1 | ···················· | ···················· | ···················· | ···················· | ···················· | ···················· |
| U | 2774 | hCoV-19/USA/MD-MDH-0466/2020\|EPI_ISL_682076\|2020-1 | ···················· | ···················· | ···················· | ···················· | ···················· | ···················· |
| U | 2775 | hCoV-19/USA/MD-MDH-0467/2020\|EPI_ISL_682077\|2020-1 | ···················· | ···················· | ···················· | ···················· | ···················· | ···················· |
| U | 2776 | hCoV-19/USA/MD-MDH-0468/2020\|EPI_ISL_682078\|2020-1 | ···················· | ···················· | ···················· | ···················· | ···················· | ···················· |
| U | 2777 | hCoV-19/USA/MI-UM-10036686754/2020\|EPI_ISL_682079\| | ···················· | ···················· | ···················· | ···················· | ···················· | ···················· |
| U | 2778 | hCoV-19/USA/MI-UM-10036700585/2020\|EPI_ISL_682080\| | ···················· | ·A·················· | ···················· | ···················· | ···················· | ···················· |
| U | 2779 | hCoV-19/USA/MI-UM-10036706124/2020\|EPI_ISL_682081\| | ···················· | ···················· | ···················· | ···················· | ···················· | ···················· |
| U | 2780 | hCoV-19/USA/MI-UM-10036690353/2020\|EPI_ISL_682082\| | ···················· | ···················· | ···················· | ···················· | ···················· | ···················· |
| U | 2781 | hCoV-19/USA/MI-UM-10036686607/2020\|EPI_ISL_682083\| | ···················· | ···················· | ···················· | ···················· | ···················· | ···················· |
| U | 2782 | hCoV-19/USA/MI-UM-10036686132/2020\|EPI_ISL_682084\| | ···················· | ···················· | ···················· | ···················· | ···················· | ···················· |
| U | 2783 | hCoV-19/USA/MI-UM-10036691355/2020\|EPI_ISL_682085\| | ···················· | ···················· | ···················· | ···················· | ···················· | ···················· |
| U | 2786 | hCoV-19/USA/MI-UM-10036681789/2020\|EPI_ISL_682088\| | ···················· | ·A·················· | ···················· | ···················· | ···················· | ···················· |
| U | 2787 | hCoV-19/USA/MI-UM-10036684769/2020\|EPI_ISL_682089\| | ···················· | ·············A······ | ···················· | ···················· | ···················· | ···················· |
| U | 2788 | hCoV-19/USA/MI-UM-10036680667/2020\|EPI_ISL_682090\| | ···················· | ···················· | ···················· | ···················· | ···················· | ···················· |
| U | 2789 | hCoV-19/USA/MI-UM-10036690391/2020\|EPI_ISL_682091\| | ···················· | ···················· | ···················· | ···················· | ···················· | ···················· |
| U | 2790 | hCoV-19/USA/MI-UM-10036689177/2020\|EPI_ISL_682092\| | ···················· | ···················· | ···················· | ···················· | ···················· | ···················· |
| U | 2791 | hCoV-19/USA/MI-UM-10036692128/2020\|EPI_ISL_682093\| | ···················· | ···················· | ···················· | ···················· | ···················· | ···················· |
| U | 2792 | hCoV-19/USA/MI-UM-10036701487/2020\|EPI_ISL_682094\| | ···················· | ·A·················· | ···················· | ···················· | ···················· | ···················· |
| U | 2793 | hCoV-19/USA/MI-UM-10036709269/2020\|EPI_ISL_682095\| | ···················· | ···················· | ···················· | ···················· | ···················· | ···················· |
| U | 2794 | hCoV-19/USA/MI-UM-10036688934/2020\|EPI_ISL_682097\| | ···················· | ···················· | ···················· | ···················· | ···················· | ···················· |
| U | 2795 | hCoV-19/USA/MI-UM-10036686909/2020\|EPI_ISL_682098\| | ···················· | ···················· | ···················· | ···················· | ···················· | ···················· |
| U | 2796 | hCoV-19/USA/MI-UM-10036686737/2020\|EPI_ISL_682099\| | ···················· | ···················· | ···················· | ···················· | ···················· | ···················· |
| U | 2798 | hCoV-19/mink/Denmark/mDK-571/2020\|EPI_ISL_683307\|2 | ···················· | ·A·················· | ···················· | ···················· | ···················· | ···················· |
| U | 2799 | hCoV-19/mink/Denmark/mDK-575/2020\|EPI_ISL_683309\|2 | ···················· | ·A·················· | ···················· | ···················· | ···················· | ···················· |
| U | 2800 | hCoV-19/mink/Denmark/mDK-585/2020\|EPI_ISL_683319\|2 | ···················· | ·A·················· | ···················· | ···················· | ···················· | ···················· |
| U | 2801 | hCoV-19/mink/Denmark/mDK-586/2020\|EPI_ISL_683320\|2 | ···················· | ·A·················· | ···················· | ···················· | ···················· | ···················· |
| U | 2802 | hCoV-19/mink/Denmark/mDK-587/2020\|EPI_ISL_683321\|2 | ···················· | ·A·················· | ···················· | ···················· | ···················· | ···················· |
| U | 2803 | hCoV-19/Tunisia/4107/2020\|EPI_ISL_683329\|2020-03-1 | ···················· | ···················· | ···················· | ···················· | ···················· | ···················· |
| U | 2804 | hCoV-19/France/ARA-SC011/2020\|EPI_ISL_683331\|2020- | ···················· | ···················· | ···················· | ···················· | ···················· | ···················· |
| U | 2805 | hCoV-19/France/ARA-SC032/2020\|EPI_ISL_683332\|2020- | ···················· | ···················· | ···················· | ···················· | ···················· | ···················· |
| U | 2806 | hCoV-19/France/ARA-101012/2020\|EPI_ISL_683335\|2020 | ···················· | ···················· | ···················· | ···················· | ···················· | ···················· |
| U | 2807 | hCoV-19/France/ARA-101082/2020\|EPI_ISL_683336\|2020 | ···················· | ···················· | ···················· | ···················· | ···················· | ···················· |
| U | 2808 | hCoV-19/France/ARA-101286/2020\|EPI_ISL_683337\|2020 | ···················· | ···················· | ···················· | ···················· | ···················· | ···················· |
| U | 2809 | hCoV-19/France/ARA-101325/2020\|EPI_ISL_683338\|2020 | ···················· | ···················· | ···················· | ···················· | ···················· | ···················· |
| U | 2810 | hCoV-19/France/ARA-101389/2020\|EPI_ISL_683339\|2020 | ···················· | ···················· | ···················· | ···················· | ···················· | ···················· |
| U | 2811 | hCoV-19/France/ARA-102608/2020\|EPI_ISL_683340\|2020 | ···················· | ···················· | ···················· | ···················· | ···················· | ···················· |
| U | 2812 | hCoV-19/France/ARA-102965/2020\|EPI_ISL_683341\|2020 | ···················· | ···················· | ···················· | ···················· | ···················· | ···················· |
| U | 2813 | hCoV-19/France/ARA-103182/2020\|EPI_ISL_683342\|2020 | ···················· | ···················· | ···················· | ···················· | ···················· | ···················· |
| U | 2814 | hCoV-19/France/ARA-103382/2020\|EPI_ISL_683343\|2020 | ···················· | ···················· | ···················· | ···················· | ···················· | ···················· |
| U | 2815 | hCoV-19/France/ARA-68082/2020\|EPI_ISL_683396\|2020- | ···················· | ···················· | ···················· | ···················· | ···················· | ···················· |
| U | 2816 | hCoV-19/France/ARA-75260/2020\|EPI_ISL_683399\|2020- | ···················· | ···················· | ···················· | ···················· | ···················· | ···················· |
| U | 2817 | hCoV-19/England/MILK-B8D370/2020\|EPI_ISL_673905\|20 | ···················· | ···················· | ···················· | ···················· | ···················· | ···················· |
| U | 2818 | hCoV-19/England/MILK-B8E676/2020\|EPI_ISL_673906\|20 | ···················· | ···················· | ···················· | ···················· | ···················· | ···················· |
| U | 2819 | hCoV-19/England/MILK-B8E56A/2020\|EPI_ISL_673907\|20 | ···················· | ···················· | ···················· | ···················· | ···················· | ···················· |
| U | 2820 | hCoV-19/England/MILK-B85DF2/2020\|EPI_ISL_673908\|20 | ···················· | ···················· | ···················· | ···················· | ···················· | ···················· |
| U | 2821 | hCoV-19/England/MILK-B8E588/2020\|EPI_ISL_673909\|20 | ···················· | ···················· | ···················· | ···················· | ···················· | ···················· |
| U | 2822 | hCoV-19/England/MILK-B864ED/2020\|EPI_ISL_673910\|20 | ···················· | ···················· | ···················· | ···················· | ···················· | ···················· |
| U | 2823 | hCoV-19/England/MILK-B862B6/2020\|EPI_ISL_673911\|20 | ···················· | ···················· | ···················· | ···················· | ···················· | ···················· |
| U | 2824 | hCoV-19/England/MILK-B86395/2020\|EPI_ISL_673912\|20 | ···················· | ···················· | ···················· | ···················· | ···················· | ···················· |
| U | 2825 | hCoV-19/England/MILK-B862F2/2020\|EPI_ISL_673913\|20 | ···················· | ···················· | ···················· | ···················· | ···················· | ···················· |
| U | 2826 | hCoV-19/England/MILK-B8666F/2020\|EPI_ISL_673914\|20 | ···················· | ···················· | ···················· | ···················· | ···················· | ···················· |
| U | 2827 | hCoV-19/England/MILK-B8E685/2020\|EPI_ISL_673916\|20 | ···················· | ···················· | ···················· | ···················· | ···················· | ···················· |
| U | 2828 | hCoV-19/England/MILK-B8E834/2020\|EPI_ISL_673917\|20 | ···················· | ···················· | ···················· | ···················· | ···················· | ···················· |
| U | 2829 | hCoV-19/England/MILK-B86878/2020\|EPI_ISL_673919\|20 | ···················· | ···················· | ···················· | ···················· | ···················· | ···················· |
| U | 2830 | hCoV-19/England/MILK-B8D39E/2020\|EPI_ISL_673920\|20 | ···················· | ···················· | ···················· | ···················· | ···················· | ···················· |
| U | 2831 | hCoV-19/England/MILK-B85D98/2020\|EPI_ISL_673921\|20 | ···················· | ···················· | ···················· | ···················· | ···················· | ···················· |
| U | 2832 | hCoV-19/England/MILK-B86025/2020\|EPI_ISL_673922\|20 | ···················· | ···················· | ···················· | ···················· | ···················· | ···················· |
| U | 2833 | hCoV-19/England/MILK-B85B9E/2020\|EPI_ISL_673923\|20 | ···················· | ···················· | ···················· | ···················· | ······T············· | ···················· |
| U | 2834 | hCoV-19/England/MILK-B8E500/2020\|EPI_ISL_673924\|20 | ···················· | ···················· | ···················· | ···················· | ···················· | ···················· |
| U | 2835 | hCoV-19/England/MILK-B85F92/2020\|EPI_ISL_673925\|20 | ···················· | ···················· | ···················· | ···················· | ···················· | ···················· |
| U | 2836 | hCoV-19/England/MILK-B85EA4/2020\|EPI_ISL_673927\|20 | ···················· | ···················· | ···················· | ···················· | ···················· | ···················· |
| U | 2837 | hCoV-19/England/MILK-B8D61D/2020\|EPI_ISL_673929\|20 | ···················· | ···················· | ···················· | ···················· | ···················· | ···················· |
| U | 2838 | hCoV-19/England/MILK-B85E86/2020\|EPI_ISL_673930\|20 | ···················· | ···················· | ···················· | ···················· | ···················· | ···················· |
| U | 2839 | hCoV-19/England/MILK-B860AD/2020\|EPI_ISL_673931\|20 | ···················· | ·A·················· | ···················· | ···················· | ···················· | ···A················ |
| U | 2840 | hCoV-19/England/MILK-B8E7BF/2020\|EPI_ISL_673932\|20 | ···················· | ···················· | ···················· | ···················· | ···················· | ···················· |
| U | 2841 | hCoV-19/England/MILK-B8626B/2020\|EPI_ISL_673934\|20 | ···················· | ···················· | ···················· | ···················· | ···················· | ···················· |
| U | 2842 | hCoV-19/England/MILK-B8690C/2020\|EPI_ISL_673935\|20 | ···················· | ···················· | ···················· | ···················· | ···················· | ···················· |
| U | 2843 | hCoV-19/England/MILK-B8659F/2020\|EPI_ISL_673936\|20 | ···················· | ···················· | ···················· | ···················· | ···················· | ···················· |
| U | 2844 | hCoV-19/England/MILK-B8E5E2/2020\|EPI_ISL_673937\|20 | ···················· | ···················· | ···················· | ···················· | ···················· | ···················· |
| U | 2845 | hCoV-19/England/MILK-B8D4E6/2020\|EPI_ISL_673938\|20 | ···················· | ···················· | ···················· | ···················· | ···················· | ···················· |
| U | 2846 | hCoV-19/England/MILK-B862E3/2020\|EPI_ISL_673939\|20 | ···················· | ·A·················· | ···················· | ···················· | ···················· | ···A················ |
| U | 2847 | hCoV-19/England/MILK-B96013/2020\|EPI_ISL_676100\|20 | ···················· | ·A·················· | ···················· | ···················· | ···················· | ···A················ |
| U | 2848 | hCoV-19/England/MILK-B966A8/2020\|EPI_ISL_676101\|20 | ···················· | ···················· | ···················· | ···················· | ······T············· | ···················· |
| U | 2849 | hCoV-19/England/MILK-B94A71/2020\|EPI_ISL_676102\|20 | ···················· | ···················· | ···················· | ···················· | ······T············· | ···················· |
| U | 2850 | hCoV-19/England/MILK-B93993/2020\|EPI_ISL_676103\|20 | ···················· | ···················· | ···················· | ···················· | ···················· | ···················· |
| U | 2851 | hCoV-19/England/MILK-B96893/2020\|EPI_ISL_676104\|20 | ···················· | ···················· | ···················· | ···················· | ···················· | ···················· |
| U | 2852 | hCoV-19/England/MILK-B93BE8/2020\|EPI_ISL_676106\|20 | ···················· | ···················· | ···················· | ···················· | ···················· | ···················· |
| U | 2853 | hCoV-19/England/MILK-B93C03/2020\|EPI_ISL_676107\|20 | ···················· | ···················· | ···················· | ···················· | ···················· | ···················· |
| U | 2854 | hCoV-19/England/MILK-B95FBD/2020\|EPI_ISL_676108\|20 | ···················· | ···················· | ···················· | ···················· | ···················· | ···················· |

```
U    2855 hCoV-19/England/MILK-B966F3/2020|EPI_ISL_676109|20     ····················  ····················  ·····················  ····················  ····················  ····················
U    2856 hCoV-19/England/MILK-B94F09/2020|EPI_ISL_676110|20     ····················  ····················  ·····················  ····················  ····················  ····················
U    2857 hCoV-19/England/MILK-B96A9D/2020|EPI_ISL_676111|20     ····················  ····················  ·····················  ····················  ····················  ····················
U    2858 hCoV-19/England/MILK-B94886/2020|EPI_ISL_676113|20     ····················  ····················  ·····················  ····················  ······T·············  ····················
U    2859 hCoV-19/England/MILK-B94ADB/2020|EPI_ISL_676115|20     ····················  ······A·············  ·····················  ····················  ····················  ····················
U    2860 hCoV-19/England/MILK-B96884/2020|EPI_ISL_676116|20     ····················  ····················  ·····················  ····················  ····················  ····················
U    2861 hCoV-19/England/MILK-B96004/2020|EPI_ISL_676117|20     ····················  ····················  ·····················  ····················  ····················  ····················
U    2862 hCoV-19/England/MILK-B94877/2020|EPI_ISL_676118|20     ····················  ····················  ·····················  ····················  ····················  ····················
U    2863 hCoV-19/England/MILK-B93D79/2020|EPI_ISL_676119|20     ·T··················  ····················  ·····················  ····················  ····················  ····················
U    2864 hCoV-19/England/MILK-B96365/2020|EPI_ISL_676121|20     ····················  ····A···············  ·····················  ····················  ····················  ····················
U    2865 hCoV-19/England/MILK-B939DF/2020|EPI_ISL_676122|20     ····················  ····················  ·····················  ····················  ····················  ····················
U    2866 hCoV-19/England/MILK-B9475C/2020|EPI_ISL_676124|20     ····················  ····················  ·····················  ····················  ····················  ····················
U    2867 hCoV-19/England/MILK-B93939/2020|EPI_ISL_676125|20     ····················  ····················  ·····················  ····················  ····················  ····················
U    2868 hCoV-19/England/MILK-B967A5/2020|EPI_ISL_676126|20     ····················  ····················  ·····················  ····················  ····················  ····················
U    2869 hCoV-19/England/MILK-B9430D/2020|EPI_ISL_676127|20     ····················  ····················  ·····················  ····················  ····················  ····················
U    2870 hCoV-19/England/MILK-B93D4C/2020|EPI_ISL_676129|20     ····················  ····················  ·····················  ····················  ····················  ····················
U    2871 hCoV-19/England/MILK-B93DD3/2020|EPI_ISL_676130|20     ····················  ····················  ·····················  ····················  ····················  ····················
U    2872 hCoV-19/England/MILK-B9659C/2020|EPI_ISL_676131|20     ····················  ····················  ·····················  ····················  ····················  ····················
U    2873 hCoV-19/England/MILK-B95F17/2020|EPI_ISL_676132|20     ····················  ·A··················  ·····················  ····················  ····················  ··A·················
U    2874 hCoV-19/England/MILK-B95DFF/2020|EPI_ISL_676133|20     ····················  ····················  ·····················  ····················  ····················  ····················
U    2875 hCoV-19/England/MILK-B943EF/2020|EPI_ISL_676134|20     ····················  ····················  ·····················  ····················  ····················  ····················
U    2876 hCoV-19/England/MILK-B946E6/2020|EPI_ISL_676135|20     ····················  ····················  ·····················  ····················  ····················  ····················
U    2877 hCoV-19/England/MILK-B94E75/2020|EPI_ISL_676137|20     ····················  ····················  ·····················  ····················  ····················  ····················
U    2878 hCoV-19/England/MILK-B9681B/2020|EPI_ISL_676138|20     ····················  ····················  ·····················  ····················  ····················  ····················
U    2879 hCoV-19/England/MILK-B96031/2020|EPI_ISL_676180|20     ····················  ····················  ·····················  ····················  ····················  ····················
U    2880 hCoV-19/England/MILK-B96602/2020|EPI_ISL_676181|20     ····················  ····················  ·····················  ····················  ····················  ····················
U    2881 hCoV-19/England/MILK-B944DD/2020|EPI_ISL_676182|20     ····················  ····················  ·····················  ····················  ····················  ····················
U    2882 hCoV-19/England/MILK-B9426A/2020|EPI_ISL_676190|20     ····················  ····················  ·····················  ····················  ····················  ····················
U    2883 hCoV-19/England/MILK-B968FD/2020|EPI_ISL_676191|20     ····················  ····················  ·····················  ····················  ····················  ····················
U    2884 hCoV-19/England/MILK-B94112/2020|EPI_ISL_676192|20     ····················  ····················  ·····················  ····················  ····················  ····················
U    2885 hCoV-19/England/MILK-B96909/2020|EPI_ISL_676193|20     ····················  ····················  ·····················  ····················  ····················  ····················
U    2886 hCoV-19/England/MILK-B94AEA/2020|EPI_ISL_676195|20     ····················  ····················  ·····················  ····················  ····················  ····················
U    2887 hCoV-19/England/MILK-B94A80/2020|EPI_ISL_676196|20     ····················  ····················  ·····················  ····················  ····················  ····················
U    2888 hCoV-19/England/MILK-B949CF/2020|EPI_ISL_676198|20     ····················  ····················  ·····················  ····················  ····················  ····················
U    2889 hCoV-19/England/MILK-B94DB4/2020|EPI_ISL_676199|20     ····················  ····················  ·····················  ····················  ····················  ····················
U    2890 hCoV-19/USA/WI-UW-1992/2020|EPI_ISL_677400|2020-09     ····················  ····················  ·····················  ····················  ····················  ····················
U    2891 hCoV-19/USA/WI-UW-1993/2020|EPI_ISL_677401|2020-09     ····················  ····················  ·····················  ····················  ····················  ····················
U    2892 hCoV-19/USA/WI-UW-1994/2020|EPI_ISL_677402|2020-09     ····················  ····················  ·····················  ····················  ····················  ····················
U    2893 hCoV-19/USA/WI-UW-1995/2020|EPI_ISL_677403|2020-09     ····················  ····················  ·····················  ····················  ····················  ····················
U    2894 hCoV-19/USA/WI-UW-1996/2020|EPI_ISL_677404|2020-09     ····················  ····················  ·····················  ····················  ····················  ····················
U    2895 hCoV-19/USA/WI-UW-1997/2020|EPI_ISL_677405|2020-09     ····················  ····················  ·····················  ····················  ····················  ····················
U    2896 hCoV-19/USA/WI-UW-1998/2020|EPI_ISL_677406|2020-09     ····················  ····················  ·····················  ····················  ····················  ····················
U    2897 hCoV-19/USA/WI-UW-1999/2020|EPI_ISL_677407|2020-09     ····················  ····················  ·····················  ····················  ····················  ····················
U    2898 hCoV-19/USA/WI-UW-2000/2020|EPI_ISL_677408|2020-09     ····················  ····················  ·····················  ····················  ····················  ····················
U    2899 hCoV-19/USA/WI-UW-2001/2020|EPI_ISL_677409|2020-09     ····················  ·A··················  ·····················  ····················  ····················  ····················
U    2900 hCoV-19/USA/WI-UW-2002/2020|EPI_ISL_677410|2020-09     ····················  ····················  ·····················  ····················  ····················  ····················
U    2901 hCoV-19/USA/WI-UW-2003/2020|EPI_ISL_677411|2020-09     ····················  ····················  ·····················  ····················  ····················  ····················
U    2902 hCoV-19/USA/WI-UW-2004/2020|EPI_ISL_677412|2020-09     ····················  ····················  ·····················  ····················  ····················  ····················
U    2903 hCoV-19/USA/WI-UW-2005/2020|EPI_ISL_677413|2020-09     ····················  ····················  ·····················  ····················  ····················  ····················
U    2904 hCoV-19/USA/WI-UW-2006/2020|EPI_ISL_677414|2020-09     ····················  ····················  ·····················  ····················  ····················  ····················
U    2905 hCoV-19/USA/WI-UW-2007/2020|EPI_ISL_677415|2020-09     ····················  ····················  ·····················  ····················  ····················  ····················
U    2906 hCoV-19/USA/WI-UW-2008/2020|EPI_ISL_677416|2020-09     ····················  ·A··················  ·····················  ····················  ····················  ····················
U    2907 hCoV-19/USA/WI-UW-2009/2020|EPI_ISL_677417|2020-09     ····················  ····················  ·····················  ····················  ····················  ····················
U    2908 hCoV-19/USA/WI-UW-2010/2020|EPI_ISL_677418|2020-09     ····················  ····················  ·····················  ····················  ····················  ····················
U    2909 hCoV-19/USA/WI-UW-2011/2020|EPI_ISL_677419|2020-09     ····················  ····················  ·····················  ····················  ····················  ····················
U    2910 hCoV-19/USA/WI-UW-2012/2020|EPI_ISL_677420|2020-09     ····················  ····················  ·····················  ····················  ····················  ····················
U    2911 hCoV-19/USA/WI-UW-2014/2020|EPI_ISL_677421|2020-09     ····················  ····················  ·····················  ····················  ····················  ····················
U    2912 hCoV-19/USA/WI-UW-2015/2020|EPI_ISL_677422|2020-10     ····················  ····················  ·····················  ····················  ····················  ····················
U    2930 hCoV-19/USA/WI-UW-2034/2020|EPI_ISL_677440|2020-09     ····················  ····················  ·····················  ····················  ····················  ····················
U    2943 hCoV-19/USA/WI-UW-2050/2020|EPI_ISL_677455|2020-10     ····················  ····················  ·····················  ····················  ····················  ····················
U    2944 hCoV-19/USA/WI-UW-2051/2020|EPI_ISL_677456|2020-10     ····················  ····················  ·····················  ····················  ····················  ····················
U    2946 hCoV-19/USA/IL-UW-2053/2020|EPI_ISL_677458|2020-10     ····················  ····················  ·····················  ····················  ····················  ····················
U    2947 hCoV-19/USA/WI-UW-2054/2020|EPI_ISL_677459|2020-10     ····················  ····················  ·····················  ····················  ····················  ····················
U    2948 hCoV-19/USA/WI-UW-2055/2020|EPI_ISL_677460|2020-10     ····················  ····················  ·····················  ····················  ····················  ····················
U    2949 hCoV-19/USA/WI-UW-2056/2020|EPI_ISL_677461|2020-10     ····················  ····················  ·····················  ····················  ····················  ····················
U    2951 hCoV-19/USA/WI-UW-2058/2020|EPI_ISL_677463|2020-10     ····················  ····················  ·····················  ····················  ····················  ····················
U    2955 hCoV-19/USA/WI-UW-2062/2020|EPI_ISL_677467|2020-07     ····················  ····················  ·····················  ····················  ····················  ····················
U    2956 hCoV-19/USA/IL-UW-2063/2020|EPI_ISL_677468|2020-10     ····················  ····················  ·····················  ····················  ····················  ····················
U    2957 hCoV-19/USA/WI-UW-2064/2020|EPI_ISL_677469|2020-10     ····················  ····················  ·····················  ····················  ····················  ····················
U    2958 hCoV-19/England/204700725/2020|EPI_ISL_678713|2020     ····················  ····················  ·····················  ····················  ····················  ····················
U    2959 hCoV-19/England/204700726/2020|EPI_ISL_678714|2020     ····················  ····················  ·····················  ····················  ······T·············  ····················
U    2960 hCoV-19/England/204720526/2020|EPI_ISL_678716|2020     ····················  ····················  ·····················  ····················  ····················  ····················
U    2961 hCoV-19/England/204720532/2020|EPI_ISL_678717|2020     ····················  ····················  ·····················  ····················  ······T·············  ····················
U    2962 hCoV-19/England/204720536/2020|EPI_ISL_678719|2020     ····················  ····················  ·····················  ····················  ····················  ····················
U    2963 hCoV-19/England/204720537/2020|EPI_ISL_678720|2020     ····················  ····················  ·····················  ····················  ····················  ····················
U    2964 hCoV-19/England/204721103/2020|EPI_ISL_678722|2020     ····················  ····················  ·····················  ····················  ····················  ····················
U    2965 hCoV-19/England/204721246/2020|EPI_ISL_678723|2020     ····················  ····················  ·····················  ····················  ····················  ····················
U    2966 hCoV-19/England/204721277/2020|EPI_ISL_678724|2020     ····················  ····················  ·····················  ····················  ····················  ····················
U    2967 hCoV-19/England/204742309/2020|EPI_ISL_678730|2020     ····················  ····················  ·····················  ····················  ····················  ····················
U    2968 hCoV-19/England/204761558/2020|EPI_ISL_678738|2020     ····················  ····················  ·····················  ····················  ······T·············  ····················
```

| | | | | | | | | |
|---|---|---|---|---|---|---|---|---|
| U | 2969 | hCoV-19/England/204780727/2020\|EPI_ISL_678745\|2020 | ···················· | ···················· | ···················· | ···················· | ···················· | ···················· |
| U | 2970 | hCoV-19/England/204821548/2020\|EPI_ISL_678775\|2020 | ···················· | ···················· | ···················· | ···················· | ···················· | ···················· |
| U | 2971 | hCoV-19/England/204821612/2020\|EPI_ISL_678776\|2020 | ···················· | ···················· | ···················· | ···················· | ···················· | ···················· |
| U | 2972 | hCoV-19/England/204821908/2020\|EPI_ISL_678778\|2020 | ···················· | ···················· | ···················· | ···················· | ···················· | ···················· |
| U | 2973 | hCoV-19/England/204841810/2020\|EPI_ISL_678780\|2020 | ···················· | ···················· | ···················· | ···················· | ···················· | ···················· |
| U | 2974 | hCoV-19/USA/PA-MGEL-00412/2020\|EPI_ISL_682013\|2020 | ···················· | ···················· | ···················· | ···················· | ···················· | ···················· |
| U | 2975 | hCoV-19/USA/PA-MGEL-00413/2020\|EPI_ISL_682014\|2020 | ···················· | ···················· | ···················· | ···················· | ···················· | ···················· |
| U | 2976 | hCoV-19/USA/PA-MGEL-00417/2020\|EPI_ISL_682015\|2020 | ···················· | ···················· | ···················· | ···················· | ···················· | ···················· |
| U | 2977 | hCoV-19/USA/PA-MGEL-00418/2020\|EPI_ISL_682016\|2020 | ···················· | ···················· | ···················· | ···················· | ···················· | ···················· |
| U | 2978 | hCoV-19/USA/PA-MGEL-00420/2020\|EPI_ISL_682017\|2020 | ···················· | ···················· | ···················· | ···················· | ···················· | ···················· |
| U | 2979 | hCoV-19/USA/PA-MGEL-00422/2020\|EPI_ISL_682018\|2020 | ···················· | ···················· | ···················· | ···················· | ···················· | ···················· |
| U | 2980 | hCoV-19/USA/PA-MGEL-00436/2020\|EPI_ISL_682019\|2020 | ···················· | ···················· | ···················· | ···················· | ···················· | ···················· |
| U | 2981 | hCoV-19/USA/PA-MGEL-00439/2020\|EPI_ISL_682020\|2020 | ···················· | ···················· | ···················· | ···················· | ···················· | ···················· |
| U | 2982 | hCoV-19/USA/PA-MGEL-00440/2020\|EPI_ISL_682021\|2020 | ···················· | ···················· | ···················· | ···················· | ···················· | ···················· |
| U | 2983 | hCoV-19/USA/PA-MGEL-00441/2020\|EPI_ISL_682022\|2020 | ···················· | ···················· | ···················· | ···················· | ···················· | ···················· |
| U | 2984 | hCoV-19/USA/PA-MGEL-00473/2020\|EPI_ISL_682023\|2020 | ···················· | ···················· | ···················· | ···················· | ···················· | ···················· |
| U | 2985 | hCoV-19/USA/PA-MGEL-00475/2020\|EPI_ISL_682024\|2020 | ···················· | ···················· | ···················· | ···················· | ···················· | ···················· |
| U | 2986 | hCoV-19/USA/PA-MGEL-00486/2020\|EPI_ISL_682025\|2020 | ···················· | ···················· | ···················· | ···················· | ···················· | ···················· |
| U | 2987 | hCoV-19/USA/PA-MGEL-00500/2020\|EPI_ISL_682026\|2020 | ···················· | ···················· | ···················· | ···················· | ···················· | ···················· |
| U | 2988 | hCoV-19/USA/PA-MGEL-00501/2020\|EPI_ISL_682027\|2020 | ···················· | ···················· | ···················· | ···················· | ···················· | ···················· |
| U | 2989 | hCoV-19/USA/PA-MGEL-00502/2020\|EPI_ISL_682028\|2020 | ···················· | ···················· | ···················· | ···················· | ···················· | ···················· |
| U | 2990 | hCoV-19/USA/PA-MGEL-00640/2020\|EPI_ISL_682029\|2020 | ···················· | ···················· | ···················· | ···················· | ···················· | ···················· |
| U | 2991 | hCoV-19/USA/PA-MGEL-00695/2020\|EPI_ISL_682030\|2020 | ···················· | ·A·················· | ···················· | ···················· | ···················· | ···················· |
| U | 2992 | hCoV-19/USA/PA-MGEL-00700/2020\|EPI_ISL_682031\|2020 | ···················· | ·A·················· | ···················· | ···················· | ···················· | ···················· |
| U | 2993 | hCoV-19/USA/PA-MGEL-00701/2020\|EPI_ISL_682032\|2020 | ···················· | ·A·················· | ···················· | ···················· | ···················· | ···················· |
| U | 2994 | hCoV-19/USA/PA-MGEL-00702/2020\|EPI_ISL_682033\|2020 | ···················· | ·A·················· | ···················· | ···················· | ···················· | ···················· |
| U | 2995 | hCoV-19/USA/PA-MGEL-00703/2020\|EPI_ISL_682034\|2020 | ···················· | ·A·················· | ···················· | ···················· | ···················· | ···················· |
| U | 2996 | hCoV-19/USA/PA-MGEL-00705/2020\|EPI_ISL_682035\|2020 | ···················· | ·A·················· | ···················· | ···················· | ···················· | ···················· |
| U | 2997 | hCoV-19/USA/PA-MGEL-00707/2020\|EPI_ISL_682036\|2020 | ···················· | ·A·················· | ···················· | ···················· | ···················· | ···················· |
| U | 2998 | hCoV-19/USA/PA-MGEL-00709/2020\|EPI_ISL_682037\|2020 | ···················· | ·A·················· | ···················· | ···················· | ···················· | ···················· |
| U | 2999 | hCoV-19/USA/PA-MGEL-00710/2020\|EPI_ISL_682038\|2020 | ···················· | ·A·················· | ···················· | ···················· | ···················· | ···················· |
| U | 3000 | hCoV-19/USA/PA-MGEL-00712/2020\|EPI_ISL_682039\|2020 | ···················· | ·A·················· | ···················· | ···················· | ···················· | ···················· |
| U | 3001 | hCoV-19/USA/PA-MGEL-00713/2020\|EPI_ISL_682040\|2020 | ···················· | ·A·················· | ···················· | ···················· | ···················· | ···················· |
| U | 3002 | hCoV-19/USA/PA-MGEL-00714/2020\|EPI_ISL_682041\|2020 | ···················· | ·A·················· | ···················· | ···················· | ···················· | ···················· |
| U | 3003 | hCoV-19/USA/PA-MGEL-00740/2020\|EPI_ISL_682042\|2020 | ···················· | ·A·················· | ···················· | ···················· | ···················· | ···················· |
| U | 3004 | hCoV-19/USA/PA-MGEL-00741/2020\|EPI_ISL_682043\|2020 | ···················· | ···················· | ···················· | ···················· | ···················· | ···················· |
| U | 3005 | hCoV-19/USA/PA-MGEL-00748/2020\|EPI_ISL_682044\|2020 | ···················· | ·A·················· | ···················· | ···················· | ···················· | ···················· |
| U | 3006 | hCoV-19/USA/PA-MGEL-00751/2020\|EPI_ISL_682045\|2020 | ···················· | ···················· | ···················· | ···················· | ···················· | ···················· |
| U | 3007 | hCoV-19/USA/PA-MGEL-00752/2020\|EPI_ISL_682046\|2020 | ···················· | ·A·················· | ···················· | ···················· | ···················· | ···················· |
| U | 3008 | hCoV-19/USA/PA-MGEL-00753/2020\|EPI_ISL_682047\|2020 | ···················· | ·A·················· | ···················· | ···················· | ···················· | ···················· |
| U | 3009 | hCoV-19/USA/PA-MGEL-00759/2020\|EPI_ISL_682048\|2020 | ···················· | ···················· | ···················· | ···················· | ···················· | ···················· |
| U | 3010 | hCoV-19/USA/PA-MGEL-00762/2020\|EPI_ISL_682049\|2020 | ···················· | ·A·················· | ···················· | ···················· | ···················· | ···················· |
| U | 3011 | hCoV-19/USA/PA-MGEL-00763/2020\|EPI_ISL_682050\|2020 | ···················· | ···················· | ···················· | ···················· | ···················· | ···················· |
| U | 3012 | hCoV-19/USA/PA-MGEL-00764/2020\|EPI_ISL_682051\|2020 | ···················· | ···················· | ···················· | ···················· | ···················· | ···················· |
| U | 3013 | hCoV-19/USA/PA-MGEL-00765/2020\|EPI_ISL_682052\|2020 | ···················· | ···················· | ···················· | ···················· | ···················· | ···················· |
| U | 3014 | hCoV-19/USA/PA-MGEL-00767/2020\|EPI_ISL_682053\|2020 | ···················· | ···················· | ···················· | ···················· | ···················· | ···················· |
| U | 3015 | hCoV-19/USA/PA-MGEL-00768/2020\|EPI_ISL_682054\|2020 | ···················· | ···················· | ···················· | ···················· | ···················· | ···················· |
| U | 3016 | hCoV-19/USA/PA-MGEL-00769/2020\|EPI_ISL_682055\|2020 | ···················· | ·A·················· | ···················· | ···················· | ···················· | ···················· |
| U | 3017 | hCoV-19/USA/PA-MGEL-00770/2020\|EPI_ISL_682056\|2020 | ···················· | ·A·················· | ···················· | ···················· | ···················· | ···················· |
| U | 3018 | hCoV-19/Ireland/MH-NVRL-20G43965/2020\|EPI_ISL_6820 | ···················· | ···················· | ···················· | ···················· | ···················· | ···················· |
| U | 3019 | hCoV-19/USA/MD-MDH-0447/2020\|EPI_ISL_682060\|2020-1 | ···················· | ·A·················· | ···················· | ···················· | ···················· | ···················· |
| U | 3020 | hCoV-19/USA/MD-MDH-0448/2020\|EPI_ISL_682061\|2020-1 | ···················· | ···················· | ···················· | ···················· | ···················· | ···················· |
| U | 3021 | hCoV-19/USA/MD-MDH-0449/2020\|EPI_ISL_682062\|2020-1 | ···················· | ···················· | ···················· | ···················· | ···················· | ···················· |
| U | 3022 | hCoV-19/USA/MD-MDH-0450/2020\|EPI_ISL_682063\|2020-1 | ···················· | ···················· | ···················· | ···················· | ···················· | ···················· |
| U | 3023 | hCoV-19/USA/MD-MDH-0451/2020\|EPI_ISL_682064\|2020-1 | ···················· | ·A·················· | ···················· | ···················· | ···················· | ···················· |
| U | 3024 | hCoV-19/USA/MD-MDH-0453/2020\|EPI_ISL_682065\|2020-1 | ···················· | ···················· | ···················· | ···················· | ···················· | ···················· |
| U | 3025 | hCoV-19/USA/MD-MDH-0454/2020\|EPI_ISL_682066\|2020-1 | ···················· | ···················· | ···················· | ···················· | ···················· | ···················· |
| U | 3026 | hCoV-19/USA/MD-MDH-0455/2020\|EPI_ISL_682067\|2020-1 | ···················· | ···················· | ···················· | ···················· | ···················· | ···················· |
| U | 3027 | hCoV-19/France/ARA-103892/2020\|EPI_ISL_683344\|2020 | ···················· | ···················· | ···················· | ···················· | ···················· | ···················· |
| U | 3028 | hCoV-19/France/ARA-103906/2020\|EPI_ISL_683345\|2020 | ···················· | ···················· | ···················· | ···················· | ···················· | ···················· |
| U | 3029 | hCoV-19/France/ARA-104138/2020\|EPI_ISL_683347\|2020 | ···················· | ···················· | ···················· | ···················· | ···················· | ···················· |
| U | 3030 | hCoV-19/France/ARA-104202/2020\|EPI_ISL_683348\|2020 | ···················· | ···················· | ···················· | ···················· | ···················· | ···················· |
| U | 3031 | hCoV-19/France/ARA-104314/2020\|EPI_ISL_683349\|2020 | ···················· | ···················· | ···················· | ···················· | ···················· | ···················· |
| U | 3032 | hCoV-19/France/ARA-104350/2020\|EPI_ISL_683350\|2020 | ···················· | ···················· | ···················· | ···················· | ···················· | ···················· |
| U | 3033 | hCoV-19/France/ARA-38358/2020\|EPI_ISL_683352\|2020- | ···················· | ···················· | ···················· | ···················· | ···················· | ···················· |
| U | 3034 | hCoV-19/France/ARA-38548/2020\|EPI_ISL_683353\|2020- | ···················· | ···················· | ···················· | ···················· | ···················· | ···················· |
| U | 3035 | hCoV-19/France/ARA-39609/2020\|EPI_ISL_683355\|2020- | ···················· | ···················· | ···················· | ···················· | ···················· | ···················· |
| U | 3036 | hCoV-19/France/ARA-40093/2020\|EPI_ISL_683356\|2020- | ···················· | ···················· | ···················· | ···················· | ···················· | ···················· |
| U | 3037 | hCoV-19/France/ARA-40115/2020\|EPI_ISL_683357\|2020- | ···················· | ···················· | ···················· | ···················· | ···················· | ···················· |
| U | 3038 | hCoV-19/France/ARA-40331/2020\|EPI_ISL_683358\|2020- | ···················· | ···················· | ···················· | ···················· | ···················· | ···················· |
| U | 3039 | hCoV-19/France/ARA-40765/2020\|EPI_ISL_683359\|2020- | ···················· | ···················· | ···················· | ···················· | ···················· | ···················· |
| U | 3040 | hCoV-19/France/ARA-41359/2020\|EPI_ISL_683362\|2020- | ···················· | ···················· | ···················· | ···················· | ···················· | ···················· |
| U | 3041 | hCoV-19/France/ARA-43052/2020\|EPI_ISL_683363\|2020- | ···················· | ···················· | ···················· | ···················· | ···················· | ···················· |
| U | 3042 | hCoV-19/France/ARA-57645/2020\|EPI_ISL_683364\|2020- | ···················· | ···················· | ···················· | ···················· | ···················· | ···················· |
| U | 3043 | hCoV-19/France/ARA-58154/2020\|EPI_ISL_683365\|2020- | ···················· | ···················· | ···················· | ···················· | ···················· | ···················· |
| U | 3044 | hCoV-19/France/ARA-58470/2020\|EPI_ISL_683366\|2020- | ···················· | ···················· | ···················· | ···················· | ···················· | ···················· |
| U | 3045 | hCoV-19/France/ARA-59557/2020\|EPI_ISL_683367\|2020- | ···················· | ···················· | ···················· | ···················· | ···················· | ···················· |
| U | 3046 | hCoV-19/France/ARA-59593/2020\|EPI_ISL_683368\|2020- | ···················· | ···················· | ···················· | ···················· | ···················· | ···················· |
| U | 3047 | hCoV-19/France/ARA-60388/2020\|EPI_ISL_683370\|2020- | ···················· | ···················· | ···················· | ···················· | ···················· | ···················· |
| U | 3048 | hCoV-19/France/ARA-60533/2020\|EPI_ISL_683371\|2020- | ···················· | ···················· | ···················· | ···················· | ···················· | ···················· |

| | | | | | | | | |
|---|---|---|---|---|---|---|---|---|
| U | 3049 | hCoV-19/France/ARA-60758/2020\|EPI_ISL_683373\|2020- | ···················· | ···················· | ···················· | ···················· | ···················· | ···················· |
| U | 3050 | hCoV-19/France/ARA-61882/2020\|EPI_ISL_683374\|2020- | ···················· | ···················· | ···················· | ···················· | ···················· | ···················· |
| U | 3051 | hCoV-19/France/ARA-62546/2020\|EPI_ISL_683375\|2020- | ···················· | ·A·················· | ···················· | ···················· | ···················· | ···················· |
| U | 3052 | hCoV-19/France/ARA-62614/2020\|EPI_ISL_683376\|2020- | ···················· | ···················· | ···················· | ···················· | ···················· | ···················· |
| U | 3053 | hCoV-19/France/ARA-62672/2020\|EPI_ISL_683377\|2020- | ···················· | ···················· | ···················· | ···················· | ···················· | ···················· |
| U | 3054 | hCoV-19/France/ARA-63944/2020\|EPI_ISL_683378\|2020- | ···················· | ···················· | ···················· | ···················· | ···················· | ···················· |
| U | 3055 | hCoV-19/France/ARA-64054/2020\|EPI_ISL_683379\|2020- | ···················· | ···················· | ···················· | ···················· | ···················· | ···················· |
| U | 3056 | hCoV-19/France/ARA-64293/2020\|EPI_ISL_683380\|2020- | ···················· | ···················· | ···················· | ···················· | ···················· | ···················· |
| U | 3057 | hCoV-19/France/ARA-64317/2020\|EPI_ISL_683381\|2020- | ···················· | ···················· | ···················· | ···················· | ···················· | ···················· |
| U | 3058 | hCoV-19/France/ARA-65734/2020\|EPI_ISL_683382\|2020- | ···················· | ···················· | ···················· | ···················· | ···················· | ···················· |
| U | 3059 | hCoV-19/France/ARA-6618/2020\|EPI_ISL_683383\|2020-0 | ···················· | ·A·················· | ···················· | ···················· | ···················· | ···················· |
| U | 3060 | hCoV-19/France/ARA-66769/2020\|EPI_ISL_683384\|2020- | ···················· | ···················· | ···················· | ···················· | ···················· | ···················· |
| U | 3061 | hCoV-19/France/ARA-67363/2020\|EPI_ISL_683385\|2020- | ···················· | ···················· | ···················· | ···················· | ···················· | ···················· |
| U | 3062 | hCoV-19/France/ARA-67638/2020\|EPI_ISL_683386\|2020- | ···················· | ···················· | ···················· | ···················· | ······T············· | ···················· |
| U | 3063 | hCoV-19/France/ARA-67649/2020\|EPI_ISL_683387\|2020- | ···················· | ···················· | ···················· | ···················· | ···················· | ···················· |
| U | 3064 | hCoV-19/France/ARA-67665/2020\|EPI_ISL_683388\|2020- | ···················· | ···················· | ···················· | ···················· | ···················· | ···················· |
| U | 3065 | hCoV-19/France/ARA-67679/2020\|EPI_ISL_683389\|2020- | ···················· | ···················· | ···················· | ···················· | ···················· | ···················· |
| U | 3066 | hCoV-19/France/ARA-67810/2020\|EPI_ISL_683390\|2020- | ···················· | ···················· | ···················· | ···················· | ···················· | ···················· |
| U | 3067 | hCoV-19/France/ARA-68070/2020\|EPI_ISL_683392\|2020- | ···················· | ···················· | ···················· | ···················· | ···················· | ···················· |
| U | 3068 | hCoV-19/France/ARA-68071/2020\|EPI_ISL_683393\|2020- | ···················· | ···················· | ···················· | ···················· | ···················· | ···················· |
| U | 3069 | hCoV-19/France/ARA-68072/2020\|EPI_ISL_683394\|2020- | ···················· | ···················· | ···················· | ···················· | ···················· | ···················· |
| U | 3070 | hCoV-19/France/ARA-68073/2020\|EPI_ISL_683395\|2020- | ···················· | ···················· | ···················· | ···················· | ···················· | ···················· |
| U | 3071 | hCoV-19/France/ARA-68090/2020\|EPI_ISL_683397\|2020- | ···················· | ···················· | ···················· | ···················· | ···················· | ···················· |
| U | 3072 | hCoV-19/France/ARA-75253/2020\|EPI_ISL_683398\|2020- | ···················· | ···················· | ···················· | ···················· | ···················· | ···················· |
| U | 3073 | hCoV-19/England/MILK-B94938/2020\|EPI_ISL_673821\|20 | ···················· | ···················· | ···················· | ···················· | ···················· | ···················· |
| U | 3074 | hCoV-19/England/CAMC-B9A20A/2020\|EPI_ISL_673823\|20 | ···················· | ···················· | ···················· | ···················· | ···················· | ···················· |
| U | 3075 | hCoV-19/England/CAMC-B98A04/2020\|EPI_ISL_673826\|20 | ···················· | ···················· | ···················· | ···················· | ···················· | ···················· |
| U | 3076 | hCoV-19/England/MILK-B94C11/2020\|EPI_ISL_673828\|20 | ···················· | ···················· | ···················· | ···················· | ···················· | ···················· |
| U | 3077 | hCoV-19/England/MILK-B94C20/2020\|EPI_ISL_673829\|20 | ···················· | ···················· | ···················· | ···················· | ···················· | ···················· |
| U | 3078 | hCoV-19/England/CAMC-B9998E/2020\|EPI_ISL_673830\|20 | ···················· | ···················· | ···················· | ···················· | ···················· | ···················· |
| U | 3079 | hCoV-19/England/MILK-B952E1/2020\|EPI_ISL_673832\|20 | ···················· | ···················· | ···················· | ···················· | ······T············· | ···················· |
| U | 3080 | hCoV-19/England/MILK-B94B9C/2020\|EPI_ISL_673833\|20 | ···················· | ···················· | ···················· | ···················· | ···················· | ···················· |
| U | 3081 | hCoV-19/England/MILK-B95005/2020\|EPI_ISL_673834\|20 | ···················· | ···················· | ···················· | ···················· | ···················· | ···················· |
| U | 3082 | hCoV-19/England/MILK-B94929/2020\|EPI_ISL_673835\|20 | ···················· | ···················· | ···················· | ···················· | ···················· | ···················· |
| U | 3083 | hCoV-19/England/CAMC-B99E70/2020\|EPI_ISL_673836\|20 | ···················· | ···················· | ···················· | ···················· | ···················· | ···················· |
| U | 3084 | hCoV-19/England/CAMC-B988EC/2020\|EPI_ISL_673837\|20 | ···················· | ···················· | ···················· | ···················· | ···················· | ···················· |
| U | 3085 | hCoV-19/England/CAMC-B9858B/2020\|EPI_ISL_673838\|20 | ···················· | ···················· | ···················· | ···················· | ···················· | ···················· |
| U | 3086 | hCoV-19/England/CAMC-B9A6C2/2020\|EPI_ISL_673839\|20 | ···················· | ···················· | ···················· | ···················· | ···················· | ···················· |
| U | 3087 | hCoV-19/England/CAMC-B98767/2020\|EPI_ISL_673841\|20 | ···················· | ···················· | ···················· | ···················· | ···················· | ···················· |
| U | 3088 | hCoV-19/England/CAMC-B98828/2020\|EPI_ISL_673844\|20 | ···················· | ···················· | ···················· | ···················· | ···················· | ···················· |
| U | 3089 | hCoV-19/England/CAMC-B98BD4/2020\|EPI_ISL_673845\|20 | ···················· | ···················· | ···················· | ···················· | ···················· | ···················· |
| U | 3090 | hCoV-19/England/CAMC-B9910E/2020\|EPI_ISL_673846\|20 | ···················· | ···················· | ···················· | ···················· | ···················· | ···················· |
| U | 3091 | hCoV-19/England/CAMC-B98D56/2020\|EPI_ISL_673847\|20 | ···················· | ···················· | ···················· | ···················· | ···················· | ···················· |
| U | 3092 | hCoV-19/England/MILK-B94BC9/2020\|EPI_ISL_673848\|20 | ···················· | ···················· | ···················· | ···················· | ···················· | ···················· |
| U | 3093 | hCoV-19/England/CAMC-B9AED9/2020\|EPI_ISL_673849\|20 | ···················· | ···················· | ···················· | ···················· | ···················· | ···················· |
| U | 3094 | hCoV-19/England/CAMC-B9AEAC/2020\|EPI_ISL_673850\|20 | ···················· | ···················· | ···················· | ···················· | ···················· | ···················· |
| U | 3095 | hCoV-19/England/CAMC-B9947E/2020\|EPI_ISL_673852\|20 | ···················· | ···················· | ···················· | ···················· | ···················· | ···················· |
| U | 3096 | hCoV-19/England/CAMC-B99933/2020\|EPI_ISL_673853\|20 | ···················· | ···················· | ···················· | ···················· | ···················· | ···················· |
| U | 3097 | hCoV-19/England/CAMC-B9899E/2020\|EPI_ISL_673854\|20 | ···················· | ···················· | ···················· | ···················· | ···················· | ···················· |
| U | 3098 | hCoV-19/England/CAMC-B99F9B/2020\|EPI_ISL_673855\|20 | ···················· | ···················· | ···················· | ···················· | ···················· | ···················· |
| U | 3099 | hCoV-19/England/MILK-B951D5/2020\|EPI_ISL_673856\|20 | ···················· | ···················· | ···················· | ···················· | ···················· | ···A················ |
| U | 3100 | hCoV-19/England/MILK-B94C02/2020\|EPI_ISL_673857\|20 | ···················· | ···················· | ···················· | ···················· | ···················· | ···················· |
| U | 3101 | hCoV-19/England/MILK-B95041/2020\|EPI_ISL_673859\|20 | ···················· | ···················· | ···················· | ···················· | ···················· | ···················· |
| U | 3102 | hCoV-19/England/CAMC-B9A5B6/2020\|EPI_ISL_673861\|20 | ···················· | ···················· | ···················· | ···················· | ···················· | ···················· |
| U | 3103 | hCoV-19/England/CAMC-B9AF21/2020\|EPI_ISL_673862\|20 | ···················· | ···················· | ···················· | ···················· | ···················· | ···················· |
| U | 3104 | hCoV-19/England/CAMC-B994E7/2020\|EPI_ISL_673864\|20 | ···················· | ···················· | ···················· | ···················· | ···················· | ···················· |
| U | 3105 | hCoV-19/England/MILK-B95120/2020\|EPI_ISL_673865\|20 | ···················· | ···················· | ···················· | ···················· | ···················· | ···················· |
| U | 3106 | hCoV-19/England/MILK-B94FFA/2020\|EPI_ISL_673867\|20 | ···················· | ···················· | ···················· | ···················· | ···················· | ···················· |
| U | 3107 | hCoV-19/England/CAMC-B99159/2020\|EPI_ISL_673869\|20 | ···················· | ···················· | ···················· | ···················· | ···················· | ···················· |
| U | 3108 | hCoV-19/England/CAMC-B98B10/2020\|EPI_ISL_673870\|20 | ···················· | ·A·················· | ···················· | ···················· | ···················· | ···A················ |
| U | 3109 | hCoV-19/England/MILK-B8D3E9/2020\|EPI_ISL_673871\|20 | ···················· | ···················· | ···················· | ···················· | ···················· | ···················· |
| U | 3110 | hCoV-19/England/MILK-B85F56/2020\|EPI_ISL_673873\|20 | ···················· | ·A·················· | ···················· | ···················· | ···················· | ···A················ |
| U | 3111 | hCoV-19/England/MILK-B86377/2020\|EPI_ISL_673874\|20 | ···················· | ···················· | ···················· | ···················· | ···················· | ···················· |
| U | 3112 | hCoV-19/England/MILK-B8D10D/2020\|EPI_ISL_673875\|20 | ···················· | ···················· | ···················· | ···················· | ···················· | ···················· |
| U | 3113 | hCoV-19/England/MILK-B8608F/2020\|EPI_ISL_673876\|20 | ···················· | ···················· | ···················· | ···················· | ···················· | ···················· |
| U | 3114 | hCoV-19/England/MILK-B8D6E0/2020\|EPI_ISL_673877\|20 | ···················· | ···················· | ···················· | ···················· | ···················· | ···················· |
| U | 3115 | hCoV-19/England/MILK-B8D11C/2020\|EPI_ISL_673880\|20 | ···················· | ···················· | ···················· | ···················· | ···················· | ···················· |
| U | 3116 | hCoV-19/England/MILK-B8618C/2020\|EPI_ISL_673881\|20 | ···················· | ···················· | ···················· | ···················· | ···················· | ···················· |
| U | 3117 | hCoV-19/England/MILK-B8D4D7/2020\|EPI_ISL_673884\|20 | ···················· | ···················· | ···················· | ···················· | ···················· | ···················· |
| U | 3118 | hCoV-19/England/MILK-B8D695/2020\|EPI_ISL_673885\|20 | ···················· | ···················· | ···················· | ···················· | ···················· | ···················· |
| U | 3119 | hCoV-19/England/MILK-B8D264/2020\|EPI_ISL_673886\|20 | ···················· | ···················· | ···················· | ···················· | ···················· | ···················· |
| U | 3120 | hCoV-19/England/MILK-B86869/2020\|EPI_ISL_673887\|20 | ···················· | ···················· | ···················· | ···················· | ···················· | ···················· |
| U | 3121 | hCoV-19/England/MILK-B8625C/2020\|EPI_ISL_673893\|20 | ···················· | ···················· | ···················· | ···················· | ···················· | ···················· |
| U | 3122 | hCoV-19/England/MILK-B86368/2020\|EPI_ISL_673894\|20 | ···················· | ···················· | ···················· | ···················· | ···················· | ···················· |
| U | 3123 | hCoV-19/England/MILK-B8623E/2020\|EPI_ISL_673895\|20 | ···················· | ···················· | ···················· | ···················· | ···················· | ···················· |
| U | 3124 | hCoV-19/England/MILK-B8E6FE/2020\|EPI_ISL_673896\|20 | ···················· | ···················· | ···················· | ···················· | ···················· | ···················· |
| U | 3125 | hCoV-19/England/MILK-B8E3CA/2020\|EPI_ISL_673897\|20 | ···················· | ···················· | ···················· | ···················· | ······T············· | ···················· |
| U | 3126 | hCoV-19/England/MILK-B8D194/2020\|EPI_ISL_673898\|20 | ···················· | ···················· | ···················· | ···················· | ···················· | ···················· |
| U | 3127 | hCoV-19/England/MILK-B85EB3/2020\|EPI_ISL_673899\|20 | ···················· | ···················· | ···················· | ···················· | ···················· | ···················· |
| U | 3128 | hCoV-19/England/MILK-B8D325/2020\|EPI_ISL_673901\|20 | ···················· | ···················· | ···················· | ···················· | ···················· | ···················· |

| | | | | | | | | |
|---|---|---|---|---|---|---|---|---|
| U | 3129 | hCoV-19/England/MILK-B8D4AA/2020\|EPI_ISL_673902\|20 | ···················· | ···················· | ····················· | ···················· | ···················· | ···················· |
| U | 3130 | hCoV-19/England/MILK-B8D64A/2020\|EPI_ISL_673903\|20 | ···················· | ···················· | ····················· | ···················· | ······T············· | ···················· |
| U | 3131 | hCoV-19/England/ALDP-B84E2D/2020\|EPI_ISL_676000\|20 | ···················· | ···················· | ····················· | ···················· | ···················· | ···················· |
| U | 3132 | hCoV-19/England/MILK-B895C9/2020\|EPI_ISL_676001\|20 | ···················· | ···················· | ····················· | ···················· | ···················· | ···················· |
| U | 3133 | hCoV-19/England/ALDP-B85017/2020\|EPI_ISL_676002\|20 | ···················· | ···················· | ····················· | ···················· | ···················· | ···················· |
| U | 3134 | hCoV-19/England/MILK-B894DB/2020\|EPI_ISL_676003\|20 | ···················· | ···················· | ····················· | ···················· | ···················· | ···················· |
| U | 3135 | hCoV-19/England/MILK-B8C8CD/2020\|EPI_ISL_676004\|20 | ···················· | ··············C····· | ····················· | ···················· | ···················· | ···················· |
| U | 3136 | hCoV-19/England/MILK-B89A15/2020\|EPI_ISL_676005\|20 | ···················· | ···················· | ····················· | ···················· | ···················· | ···················· |
| U | 3137 | hCoV-19/England/ALDP-B84ED2/2020\|EPI_ISL_676006\|20 | ···················· | ···················· | ····················· | ···················· | ···················· | ···················· |
| U | 3138 | hCoV-19/England/MILK-B88973/2020\|EPI_ISL_676007\|20 | ···················· | ···················· | ····················· | ···················· | ···················· | ···················· |
| U | 3139 | hCoV-19/England/MILK-B88CF2/2020\|EPI_ISL_676008\|20 | ···················· | ···················· | ····················· | ···················· | ···················· | ···················· |
| U | 3140 | hCoV-19/England/MILK-B88B31/2020\|EPI_ISL_676009\|20 | ···················· | ···················· | ····················· | ···················· | ······T············· | ···················· |
| U | 3141 | hCoV-19/England/ALDP-B84E4B/2020\|EPI_ISL_676010\|20 | ···················· | ···················· | ····················· | ···················· | ···················· | ···················· |
| U | 3142 | hCoV-19/England/MILK-B94CD5/2020\|EPI_ISL_676011\|20 | ···················· | ···················· | ····················· | ···················· | ······T············· | ···················· |
| U | 3143 | hCoV-19/England/MILK-B960C8/2020\|EPI_ISL_676013\|20 | ···················· | ···················· | ····················· | ···················· | ···················· | ···················· |
| U | 3144 | hCoV-19/England/MILK-B944FB/2020\|EPI_ISL_676014\|20 | ···················· | ···················· | ····················· | ···················· | ···················· | ···················· |
| U | 3145 | hCoV-19/England/MILK-B9392A/2020\|EPI_ISL_676015\|20 | ···················· | ···················· | ····················· | ···················· | ···················· | ···················· |
| U | 3146 | hCoV-19/England/MILK-B9674B/2020\|EPI_ISL_676016\|20 | ···················· | ···················· | ····················· | ···················· | ···················· | ···················· |
| U | 3147 | hCoV-19/England/MILK-B96453/2020\|EPI_ISL_676017\|20 | ···················· | ···················· | ····················· | ···················· | ···················· | ···················· |
| U | 3148 | hCoV-19/England/MILK-B96796/2020\|EPI_ISL_676139\|20 | ···················· | ···················· | ····················· | ···················· | ···················· | ···················· |
| U | 3149 | hCoV-19/England/MILK-B93D2E/2020\|EPI_ISL_676141\|20 | ···················· | ···················· | ····················· | ···················· | ···················· | ···················· |
| U | 3150 | hCoV-19/England/MILK-B94C8A/2020\|EPI_ISL_676142\|20 | ···················· | ···················· | ····················· | ···················· | ···················· | ···················· |
| U | 3151 | hCoV-19/England/MILK-B94EB1/2020\|EPI_ISL_676147\|20 | ···················· | ···················· | ····················· | ···················· | ···················· | ···················· |
| U | 3152 | hCoV-19/England/MILK-B96022/2020\|EPI_ISL_676148\|20 | ···················· | ···················· | ····················· | ···················· | ···················· | ···················· |
| U | 3153 | hCoV-19/England/MILK-B94DC3/2020\|EPI_ISL_676149\|20 | ···················· | ···················· | ····················· | ···················· | ···················· | ···················· |
| U | 3154 | hCoV-19/England/MILK-B95BE6/2020\|EPI_ISL_676150\|20 | ···················· | ·A·················· | ····················· | ···················· | ···················· | ···A················ |
| U | 3155 | hCoV-19/England/MILK-B963DE/2020\|EPI_ISL_676152\|20 | ···················· | ···················· | ····················· | ···················· | ···················· | ···················· |
| U | 3156 | hCoV-19/England/MILK-B93984/2020\|EPI_ISL_676154\|20 | ···················· | ·A·················· | ····················· | ···················· | ···················· | ···················· |
| U | 3157 | hCoV-19/England/MILK-B963A1/2020\|EPI_ISL_676156\|20 | ···················· | ···················· | ····················· | ···················· | ···················· | ···················· |
| U | 3158 | hCoV-19/England/MILK-B939EE/2020\|EPI_ISL_676157\|20 | ···················· | ···················· | ····················· | ···················· | ···················· | ···················· |
| U | 3159 | hCoV-19/England/MILK-B9663F/2020\|EPI_ISL_676158\|20 | ···················· | ···················· | ····················· | ···················· | ···················· | ···················· |
| U | 3160 | hCoV-19/England/MILK-B93B60/2020\|EPI_ISL_676159\|20 | ···················· | ···················· | ····················· | ···················· | ···················· | ···················· |
| U | 3161 | hCoV-19/England/MILK-B95D68/2020\|EPI_ISL_676161\|20 | ···················· | ···················· | ····················· | ···················· | ···················· | ···················· |
| U | 3162 | hCoV-19/England/MILK-B94C7B/2020\|EPI_ISL_676162\|20 | ···················· | ·A·················· | ····················· | ···················· | ···················· | ···A················ |
| U | 3163 | hCoV-19/England/MILK-B93C12/2020\|EPI_ISL_676163\|20 | ···················· | ···················· | ····················· | ···················· | ···················· | ···················· |
| U | 3164 | hCoV-19/England/MILK-B942B5/2020\|EPI_ISL_676164\|20 | ···················· | ···················· | ····················· | ···················· | ···················· | ···················· |
| U | 3165 | hCoV-19/England/MILK-B96329/2020\|EPI_ISL_676165\|20 | ···················· | ·A·················· | ····················· | ···················· | ···················· | ···A················ |
| U | 3166 | hCoV-19/England/MILK-B96699/2020\|EPI_ISL_676166\|20 | ···················· | ···················· | ····················· | ···················· | ···················· | ···················· |
| U | 3167 | hCoV-19/England/MILK-B94EDF/2020\|EPI_ISL_676167\|20 | ···················· | ···················· | ····················· | ···················· | ···················· | ···················· |
| U | 3168 | hCoV-19/England/MILK-B96A06/2020\|EPI_ISL_676168\|20 | ···················· | ···················· | ····················· | ···················· | ···················· | ···················· |
| U | 3169 | hCoV-19/England/MILK-B94956/2020\|EPI_ISL_676169\|20 | ···················· | ···················· | ····················· | ···················· | ···················· | ···················· |
| U | 3170 | hCoV-19/England/MILK-B965AB/2020\|EPI_ISL_676171\|20 | ···················· | ···················· | ····················· | ···················· | ···················· | ···················· |
| U | 3171 | hCoV-19/England/MILK-B95F80/2020\|EPI_ISL_676173\|20 | ···················· | ···················· | ····················· | ···················· | ···················· | ···················· |
| U | 3172 | hCoV-19/England/MILK-B95FF9/2020\|EPI_ISL_676174\|20 | ···················· | ···················· | ····················· | ···················· | ···················· | ···················· |
| U | 3173 | hCoV-19/England/MILK-B93D6A/2020\|EPI_ISL_676176\|20 | ···················· | ····A··············· | ····················· | ···················· | ···················· | ···················· |
| U | 3174 | hCoV-19/England/MILK-B94297/2020\|EPI_ISL_676178\|20 | ···················· | ···················· | ····················· | ···················· | ···················· | ···················· |
| U | 3175 | hCoV-19/England/MILK-B94E1B/2020\|EPI_ISL_676186\|20 | ···················· | ···················· | ····················· | ···················· | ···················· | ···················· |
| U | 3176 | hCoV-19/England/MILK-B9615C/2020\|EPI_ISL_676187\|20 | ···················· | ···················· | ····················· | ···················· | ······T············· | ···················· |
| U | 3177 | hCoV-19/England/MILK-B94E57/2020\|EPI_ISL_676188\|20 | ···················· | ···················· | ····················· | ···················· | ···················· | ···················· |
| U | 3178 | hCoV-19/England/MILK-B96374/2020\|EPI_ISL_676189\|20 | ···················· | ·A·················· | ····················· | ···················· | ···················· | ···A················ |
| U | 3179 | hCoV-19/USA/CO-CDPHE-2008181790/2020\|EPI_ISL_67731 | ···················· | ···················· | ····················· | ···················· | ···················· | ···················· |
| U | 3180 | hCoV-19/USA/CO-CDPHE-2009041479/2020\|EPI_ISL_67731 | ···················· | ·A·················· | ····················· | ···················· | ···················· | ···················· |
| U | 3181 | hCoV-19/USA/CO-CDPHE-2009042326/2020\|EPI_ISL_67731 | ···················· | ···················· | ····················· | ···················· | ···················· | ···················· |
| U | 3182 | hCoV-19/USA/CO-CDPHE-2009042743/2020\|EPI_ISL_67731 | ···················· | ···················· | ····················· | ···················· | ···················· | ···················· |
| U | 3183 | hCoV-19/USA/CO-CDPHE-2009151301/2020\|EPI_ISL_67731 | ···················· | ···················· | ····················· | ···················· | ···················· | ···················· |
| U | 3184 | hCoV-19/USA/CO-CDPHE-2009163495/2020\|EPI_ISL_67731 | ···················· | ···················· | ····················· | ···················· | ···················· | ···················· |
| U | 3185 | hCoV-19/USA/CO-CDPHE-2010200546/2020\|EPI_ISL_67731 | ···················· | ···················· | ····················· | ···················· | ···················· | ···················· |
| U | 3186 | hCoV-19/USA/CO-CDPHE-2011020335/2020\|EPI_ISL_67731 | ···················· | ···················· | ····················· | ···················· | ···················· | ···················· |
| U | 3188 | hCoV-19/USA/WI-UW-1912/2020\|EPI_ISL_677321\|2020-10 | ···················· | ···················· | ····················· | ···················· | ···················· | ···················· |
| U | 3200 | hCoV-19/USA/WI-UW-1924/2020\|EPI_ISL_677333\|2020-10 | ···················· | ···················· | ····················· | ···················· | ···················· | ···················· |
| U | 3202 | hCoV-19/USA/WI-UW-1926/2020\|EPI_ISL_677335\|2020-10 | ···················· | ···················· | ····················· | ···················· | ···················· | ···················· |
| U | 3203 | hCoV-19/USA/WI-UW-1927/2020\|EPI_ISL_677336\|2020-10 | ···················· | ···················· | ····················· | ···················· | ···················· | ···················· |
| U | 3204 | hCoV-19/USA/WI-UW-1929/2020\|EPI_ISL_677338\|2020-10 | ···················· | ···················· | ····················· | ···················· | ···················· | ···················· |
| U | 3205 | hCoV-19/USA/WI-UW-1930/2020\|EPI_ISL_677339\|2020-09 | ···················· | ···················· | ····················· | ···················· | ···················· | ···················· |
| U | 3207 | hCoV-19/USA/WI-UW-1932/2020\|EPI_ISL_677341\|2020-10 | ···················· | ···················· | ····················· | ···················· | ···················· | ···················· |
| U | 3208 | hCoV-19/USA/WI-UW-1933/2020\|EPI_ISL_677342\|2020-10 | ···················· | ···················· | ····················· | ···················· | ···················· | ···················· |
| U | 3209 | hCoV-19/USA/WI-UW-1934/2020\|EPI_ISL_677343\|2020-09 | ···················· | ···················· | ····················· | ···················· | ···················· | ···················· |
| U | 3210 | hCoV-19/USA/WI-UW-1935/2020\|EPI_ISL_677344\|2020-09 | ···················· | ···················· | ····················· | ···················· | ···················· | ···················· |
| U | 3211 | hCoV-19/USA/WI-UW-1936/2020\|EPI_ISL_677345\|2020-10 | ···················· | ···················· | ····················· | ···················· | ···················· | ···················· |
| U | 3213 | hCoV-19/USA/WI-UW-1938/2020\|EPI_ISL_677347\|2020-10 | ···················· | ···················· | ····················· | ···················· | ···················· | ···················· |
| U | 3214 | hCoV-19/USA/WI-UW-1939/2020\|EPI_ISL_677348\|2020-10 | ···················· | ···················· | ····················· | ···················· | ···················· | ···················· |
| U | 3215 | hCoV-19/USA/WI-UW-2065/2020\|EPI_ISL_677470\|2020-10 | ···················· | ···················· | ····················· | ···················· | ···················· | ···················· |
| U | 3216 | hCoV-19/USA/WI-UW-2066/2020\|EPI_ISL_677471\|2020-10 | ···················· | ···················· | ····················· | ···················· | ···················· | ···················· |
| U | 3217 | hCoV-19/USA/WI-UW-2067/2020\|EPI_ISL_677472\|2020-10 | ···················· | ···················· | ····················· | ···················· | ···················· | ···················· |
| U | 3218 | hCoV-19/USA/WI-UW-2068/2020\|EPI_ISL_677473\|2020-10 | ···················· | ···················· | ····················· | ···················· | ···················· | ···················· |
| U | 3220 | hCoV-19/USA/WI-UW-2070/2020\|EPI_ISL_677475\|2020-10 | ···················· | ···················· | ····················· | ···················· | ···················· | ···················· |
| U | 3222 | hCoV-19/USA/WI-UW-2072/2020\|EPI_ISL_677477\|2020-10 | ···················· | ···················· | ····················· | ···················· | ···················· | ···················· |
| U | 3223 | hCoV-19/USA/WI-UW-2073/2020\|EPI_ISL_677478\|2020-10 | ···················· | ···················· | ····················· | ···················· | ···················· | ···················· |
| U | 3224 | hCoV-19/USA/WI-UW-2074/2020\|EPI_ISL_677479\|2020-10 | ···················· | ···················· | ····················· | ···················· | ···················· | ···················· |
| U | 3225 | hCoV-19/USA/WI-UW-2075/2020\|EPI_ISL_677480\|2020-10 | ···················· | ···················· | ····················· | ···················· | ···················· | ···················· |

| | | | | | | | |
|---|---|---|---|---|---|---|---|
| U | 3228 hCoV-19/USA/WI-UW-2078/2020\|EPI_ISL_677483\|2020-10 | ···················· | ···················· | ···················· | ···················· | ···················· | ···················· |
| U | 3232 hCoV-19/USA/WI-UW-2082/2020\|EPI_ISL_677487\|2020-10 | ···················· | ···················· | ···················· | ···················· | ···················· | ···················· |
| U | 3233 hCoV-19/USA/WI-UW-2083/2020\|EPI_ISL_677488\|2020-10 | ···················· | ···················· | ···················· | ···················· | ···················· | ···················· |
| U | 3236 hCoV-19/USA/WI-UW-2086/2020\|EPI_ISL_677491\|2020-09 | ···················· | ···················· | ···················· | ···················· | ···················· | ···················· |
| U | 3237 hCoV-19/USA/WI-UW-2087/2020\|EPI_ISL_677492\|2020-09 | ···················· | ···················· | ···················· | ···················· | ···················· | ···················· |
| U | 3238 hCoV-19/USA/WI-UW-2088/2020\|EPI_ISL_677493\|2020-09 | ···················· | ···················· | ···················· | ···················· | ···················· | ···················· |
| U | 3239 hCoV-19/USA/WI-UW-2089/2020\|EPI_ISL_677494\|2020-09 | ···················· | ···················· | ···················· | ···················· | ···················· | ···················· |
| U | 3240 hCoV-19/USA/WI-UW-2090/2020\|EPI_ISL_677495\|2020-09 | ···················· | ·A·················· | ···················· | ···················· | ···················· | ···················· |
| U | 3243 hCoV-19/USA/WI-UW-2093/2020\|EPI_ISL_677498\|2020-11 | ···················· | ···················· | ···················· | ···················· | ···················· | ···················· |
| U | 3244 hCoV-19/USA/WI-UW-2094/2020\|EPI_ISL_677499\|2020-11 | ···················· | ···················· | ···················· | ···················· | ···················· | ···················· |
| U | 3245 hCoV-19/England/204521651/2020\|EPI_ISL_678644\|2020 | ···················· | ···················· | ···················· | ···················· | ···················· | ···················· |
| U | 3246 hCoV-19/England/204521765/2020\|EPI_ISL_678646\|2020 | ···················· | ···················· | ···················· | ···················· | ···················· | ···················· |
| U | 3247 hCoV-19/England/204522829/2020\|EPI_ISL_678647\|2020 | ···················· | ···················· | ···················· | ···················· | ···················· | ···················· |
| U | 3248 hCoV-19/England/204540240/2020\|EPI_ISL_678648\|2020 | ···················· | ···················· | ···················· | ···················· | ···················· | ···················· |
| U | 3249 hCoV-19/England/204540260/2020\|EPI_ISL_678649\|2020 | ···················· | ···················· | ···················· | ···················· | ···················· | ···················· |
| U | 3250 hCoV-19/England/204540536/2020\|EPI_ISL_678651\|2020 | ···················· | ···················· | ···················· | ···················· | ···················· | ···················· |
| U | 3251 hCoV-19/England/204540589/2020\|EPI_ISL_678652\|2020 | ···················· | ···················· | ···················· | ···················· | ···················· | ···················· |
| U | 3252 hCoV-19/England/204540592/2020\|EPI_ISL_678653\|2020 | ···················· | ···················· | ···················· | ···················· | ···················· | ···················· |
| U | 3253 hCoV-19/England/204540593/2020\|EPI_ISL_678654\|2020 | ···················· | ···················· | ···················· | ···················· | ···················· | ···················· |
| U | 3254 hCoV-19/England/204540597/2020\|EPI_ISL_678655\|2020 | ···················· | ···················· | ···················· | ···················· | ···················· | ···················· |
| U | 3255 hCoV-19/England/204540598/2020\|EPI_ISL_678656\|2020 | ···················· | ···················· | ···················· | ···················· | ···················· | ···················· |
| U | 3256 hCoV-19/England/204540967/2020\|EPI_ISL_678657\|2020 | ···················· | ···················· | ···················· | ···················· | ···················· | ···················· |
| U | 3257 hCoV-19/England/204540971/2020\|EPI_ISL_678658\|2020 | ···················· | ···················· | ···················· | ···················· | ···················· | ···················· |
| U | 3258 hCoV-19/England/204540972/2020\|EPI_ISL_678659\|2020 | ···················· | ···················· | ···················· | ···················· | ···················· | ···················· |
| U | 3259 hCoV-19/England/204540974/2020\|EPI_ISL_678660\|2020 | ···················· | ···················· | ···················· | ···················· | ···················· | ···················· |
| U | 3260 hCoV-19/England/204540979/2020\|EPI_ISL_678661\|2020 | ···················· | ···················· | ···················· | ···················· | ···················· | ···················· |
| U | 3261 hCoV-19/England/204540993/2020\|EPI_ISL_678663\|2020 | ···················· | ···················· | ···················· | ···················· | ···················· | ···················· |
| U | 3262 hCoV-19/England/204540994/2020\|EPI_ISL_678664\|2020 | ···················· | ···················· | ···················· | ···················· | ···················· | ···················· |
| U | 3263 hCoV-19/England/204541000/2020\|EPI_ISL_678665\|2020 | ···················· | ···················· | ···················· | ···················· | ···················· | ···················· |
| U | 3264 hCoV-19/England/204541008/2020\|EPI_ISL_678666\|2020 | ···················· | ·A·················· | ···················· | ···················· | ···················· | ··A················· |
| U | 3265 hCoV-19/England/204541010/2020\|EPI_ISL_678667\|2020 | ···················· | ···················· | ···················· | ···················· | ···················· | ···················· |
| U | 3266 hCoV-19/England/204541124/2020\|EPI_ISL_678668\|2020 | ···················· | ···················· | ···················· | ···················· | ···················· | ···················· |
| U | 3267 hCoV-19/England/204541203/2020\|EPI_ISL_678669\|2020 | ···················· | ···················· | ···················· | ···················· | ···················· | ···················· |
| U | 3268 hCoV-19/England/204541206/2020\|EPI_ISL_678670\|2020 | ···················· | ···················· | ···················· | ···················· | ···················· | ···················· |
| U | 3269 hCoV-19/England/204541209/2020\|EPI_ISL_678671\|2020 | ···················· | ···················· | ···················· | ···················· | ···················· | ···················· |
| U | 3270 hCoV-19/England/204541224/2020\|EPI_ISL_678672\|2020 | ···················· | ···················· | ···················· | ···················· | ···················· | ···················· |
| U | 3271 hCoV-19/England/204541226/2020\|EPI_ISL_678673\|2020 | ···················· | ···················· | ···················· | ···················· | ···················· | ···················· |
| U | 3272 hCoV-19/England/204560270/2020\|EPI_ISL_678674\|2020 | ···················· | ···················· | ···················· | ···················· | ···················· | ···················· |
| U | 3273 hCoV-19/England/204560280/2020\|EPI_ISL_678675\|2020 | ···················· | ···················· | ···················· | ···················· | ···················· | ···················· |
| U | 3274 hCoV-19/England/204560282/2020\|EPI_ISL_678676\|2020 | ···················· | ···················· | ···················· | ···················· | ···················· | ···················· |
| U | 3275 hCoV-19/England/204560308/2020\|EPI_ISL_678677\|2020 | ···················· | ···················· | ···················· | ···················· | ···················· | ···················· |
| U | 3276 hCoV-19/England/204560319/2020\|EPI_ISL_678678\|2020 | ···················· | ···················· | ···················· | ···················· | ···················· | ···················· |
| U | 3277 hCoV-19/England/204581657/2020\|EPI_ISL_678704\|2020 | ···················· | ···················· | ···················· | ···················· | ···················· | ···················· |
| U | 3278 hCoV-19/England/204700489/2020\|EPI_ISL_678706\|2020 | ···················· | ···················· | ···················· | ···················· | ···················· | ···················· |
| U | 3279 hCoV-19/England/204700491/2020\|EPI_ISL_678707\|2020 | ···················· | ···················· | ···················· | ···················· | ···················· | ···················· |
| U | 3280 hCoV-19/England/204700569/2020\|EPI_ISL_678708\|2020 | ···················· | ···················· | ···················· | ···················· | ···················· | ···················· |
| U | 3281 hCoV-19/England/204700596/2020\|EPI_ISL_678709\|2020 | ···················· | ···················· | ···················· | ···················· | ···················· | ···················· |
| U | 3282 hCoV-19/England/204700692/2020\|EPI_ISL_678710\|2020 | ···················· | ···················· | ···················· | ···················· | ···················· | ···················· |
| U | 3283 hCoV-19/England/204700718/2020\|EPI_ISL_678711\|2020 | ···················· | ···················· | ···················· | ···················· | ·····T·············· | ···················· |
| U | 3284 hCoV-19/England/204700722/2020\|EPI_ISL_678712\|2020 | ···················· | ···················· | ···················· | ···················· | ···················· | ···················· |
| U | 3285 hCoV-19/England/PORT-2EA037/2020\|EPI_ISL_679981\|20 | ···················· | ···················· | ···················· | ···················· | ·····T·············· | ···················· |
| U | 3286 hCoV-19/England/PORT-2EA152/2020\|EPI_ISL_679985\|20 | ···················· | ···················· | ···················· | ···················· | ·····T·············· | ···················· |
| U | 3287 hCoV-19/England/QEUH-A015E8/2020\|EPI_ISL_679987\|20 | ···················· | ···················· | ···················· | ···················· | ···················· | ···················· |
| U | 3288 hCoV-19/England/QEUH-A0165E/2020\|EPI_ISL_679993\|20 | ···················· | ···················· | ···················· | ···················· | ···················· | ···················· |
| U | 3289 hCoV-19/England/QEUH-A01894/2020\|EPI_ISL_679999\|20 | ···················· | ···················· | ···················· | ···················· | ···················· | ···················· |
| U | 3290 hCoV-19/Wales/PHWC-48EADB/2020\|EPI_ISL_680960\|2020 | ···················· | ···················· | ···················· | ···················· | ···················· | ···················· |
| U | 3294 hCoV-19/Wales/PHWC-48EB32/2020\|EPI_ISL_680966\|2020 | ···················· | ···················· | ···················· | ···················· | ···················· | ···················· |
| U | 3296 hCoV-19/Wales/PHWC-48EB50/2020\|EPI_ISL_680968\|2020 | ···················· | ···················· | ···················· | ···················· | ···················· | ···················· |
| U | 3297 hCoV-19/Wales/PHWC-48EB7E/2020\|EPI_ISL_680970\|2020 | ···················· | ···················· | ···················· | ···················· | ···················· | ···················· |
| U | 3298 hCoV-19/Wales/PHWC-48EB9C/2020\|EPI_ISL_680972\|2020 | ···················· | ···················· | ···················· | ···················· | ···················· | ···················· |
| U | 3299 hCoV-19/Wales/PHWC-48EBBA/2020\|EPI_ISL_680973\|2020 | ···················· | ···················· | ···················· | ···················· | NN·················· | ···················· |
| U | 3300 hCoV-19/Wales/PHWC-48EBC9/2020\|EPI_ISL_680974\|2020 | ···················· | ···················· | ···················· | ···················· | ···················· | ···················· |
| U | 3301 hCoV-19/Wales/PHWC-48EBD8/2020\|EPI_ISL_680975\|2020 | ···················· | ···················· | ···················· | ···················· | ···················· | ···················· |
| U | 3302 hCoV-19/Wales/PHWC-48EBE7/2020\|EPI_ISL_680976\|2020 | ···················· | ···················· | ···················· | ···················· | ···················· | ···················· |
| U | 3303 hCoV-19/Wales/PHWC-48EBF6/2020\|EPI_ISL_680977\|2020 | ···················· | ···················· | ···················· | ···················· | ···················· | ···················· |
| U | 3304 hCoV-19/Wales/PHWC-48EC02/2020\|EPI_ISL_680978\|2020 | ···················· | ···················· | ···················· | ···················· | ···················· | ···················· |
| U | 3305 hCoV-19/Wales/PHWC-48EC11/2020\|EPI_ISL_680979\|2020 | ···················· | ···················· | ···················· | ···················· | ···················· | ···················· |
| U | 3306 hCoV-19/Wales/PHWC-48EC20/2020\|EPI_ISL_680980\|2020 | ···················· | ···················· | ···················· | ···················· | ···················· | ···················· |
| U | 3307 hCoV-19/Wales/PHWC-48EC3F/2020\|EPI_ISL_680981\|2020 | ···················· | ···················· | ···················· | ···················· | ···················· | ···················· |
| U | 3308 hCoV-19/Wales/PHWC-48EC5D/2020\|EPI_ISL_680983\|2020 | ···················· | ···················· | ···················· | ···················· | ···················· | ···················· |
| U | 3309 hCoV-19/Wales/PHWC-48EC6C/2020\|EPI_ISL_680984\|2020 | ···················· | ···················· | ···················· | ···················· | ···················· | ···················· |
| U | 3310 hCoV-19/Wales/PHWC-48EC8A/2020\|EPI_ISL_680986\|2020 | ···················· | ···················· | ···················· | ···················· | ···················· | ···················· |
| U | 3311 hCoV-19/Wales/PHWC-48EC99/2020\|EPI_ISL_680987\|2020 | ···················· | ···················· | ···················· | ···················· | ···················· | ···················· |
| U | 3312 hCoV-19/Wales/PHWC-48ECA8/2020\|EPI_ISL_680988\|2020 | ···················· | ···················· | ···················· | ···················· | ···················· | ···················· |
| U | 3313 hCoV-19/Wales/PHWC-48ECB7/2020\|EPI_ISL_680989\|2020 | ···················· | ···················· | ···················· | ···················· | ···················· | ···················· |
| U | 3314 hCoV-19/mink/Denmark/mDK-463/2020\|EPI_ISL_683215\|2 | ···················· | ···················· | ···················· | ···················· | ···················· | ···················· |
| U | 3315 hCoV-19/mink/Denmark/mDK-465/2020\|EPI_ISL_683216\|2 | ···················· | ···················· | ···················· | ···················· | ···················· | ···················· |
| U | 3316 hCoV-19/mink/Denmark/mDK-466/2020\|EPI_ISL_683217\|2 | ···················· | ···················· | ···················· | ···················· | ···················· | ···················· |
| U | 3317 hCoV-19/mink/Denmark/mDK-467/2020\|EPI_ISL_683218\|2 | ···················· | ···················· | ···················· | ···················· | ···················· | ···················· |
| U | 3318 hCoV-19/mink/Denmark/mDK-497/2020\|EPI_ISL_683243\|2 | ···················· | ·A·················· | ···················· | ···················· | ···················· | ···················· |

| | | | | | | | |
|---|---|---|---|---|---|---|---|
| U | 3319 hCoV-19/mink/Denmark/mDK-501/2020\|EPI_ISL_683246\|2 | ···················· | ·A·················· | ···················· | ···················· | ···················· | ···················· |
| U | 3320 hCoV-19/mink/Denmark/mDK-502/2020\|EPI_ISL_683247\|2 | ···················· | ·A·················· | ···················· | ···················· | ···················· | ···················· |
| U | 3321 hCoV-19/mink/Denmark/mDK-503/2020\|EPI_ISL_683248\|2 | ···················· | ·A·················· | ···················· | ···················· | ···················· | ···················· |
| U | 3322 hCoV-19/mink/Denmark/mDK-528/2020\|EPI_ISL_683270\|2 | ···················· | ·A·················· | ···················· | ···················· | ···················· | ···················· |
| U | 3323 hCoV-19/mink/Denmark/mDK-530/2020\|EPI_ISL_683271\|2 | ···················· | ·A·················· | ···················· | ···················· | ···················· | ···················· |
| U | 3324 hCoV-19/mink/Denmark/mDK-531/2020\|EPI_ISL_683272\|2 | ···················· | ·A·················· | ···················· | ···················· | ···················· | ···················· |
| U | 3325 hCoV-19/mink/Denmark/mDK-532/2020\|EPI_ISL_683273\|2 | ···················· | ·N·················· | ···················· | ···················· | ···················· | ···················· |
| U | 3326 hCoV-19/mink/Denmark/mDK-541/2020\|EPI_ISL_683280\|2 | ···················· | ·A·················· | ···················· | ···················· | ···················· | ···················· |
| U | 3327 hCoV-19/mink/Denmark/mDK-562/2020\|EPI_ISL_683298\|2 | ···················· | ·A·················· | ···················· | ···················· | ···················· | ···················· |
| U | 3328 hCoV-19/mink/Denmark/mDK-563/2020\|EPI_ISL_683299\|2 | ···················· | ·A·················· | ···················· | ···················· | ···················· | ···················· |
| U | 3329 hCoV-19/England/CAMC-B98A13/2020\|EPI_ISL_673800\|20 | ···················· | ···················· | ···················· | ···················· | ···················· | ···················· |
| U | 3330 hCoV-19/England/MILK-B95278/2020\|EPI_ISL_673801\|20 | ···················· | ···················· | ···················· | ···················· | ······T············· | ···················· |
| U | 3331 hCoV-19/England/CAMC-B9AF4F/2020\|EPI_ISL_673802\|20 | ···················· | ···················· | ···················· | ···················· | ···················· | ···················· |
| U | 3332 hCoV-19/England/MILK-B952C3/2020\|EPI_ISL_673803\|20 | ···················· | ···················· | ···················· | ···················· | ···················· | ···················· |
| U | 3333 hCoV-19/England/CAMC-B99EE9/2020\|EPI_ISL_673804\|20 | ···················· | ···················· | ···················· | ···················· | ···················· | ···A················ |
| U | 3334 hCoV-19/England/CAMC-B98855/2020\|EPI_ISL_673805\|20 | ···················· | ···················· | ···················· | ···················· | ···················· | ···················· |
| U | 3335 hCoV-19/England/CAMC-B99E52/2020\|EPI_ISL_673808\|20 | ···················· | ···················· | ···················· | ···················· | ······T············· | ···················· |
| U | 3336 hCoV-19/England/CAMC-B999BB/2020\|EPI_ISL_673809\|20 | ···················· | ···················· | ···················· | ···················· | ···················· | ···················· |
| U | 3337 hCoV-19/England/MILK-B9530C/2020\|EPI_ISL_673812\|20 | ···················· | ·A·················· | ···················· | ···················· | ···················· | ···················· |
| U | 3338 hCoV-19/England/MILK-B95269/2020\|EPI_ISL_673813\|20 | ···················· | ····A··············· | ···················· | ···················· | ···················· | ···················· |
| U | 3339 hCoV-19/England/MILK-B95023/2020\|EPI_ISL_673814\|20 | ···················· | ·A·················· | ···················· | ···················· | ···················· | ···A················ |
| U | 3340 hCoV-19/England/CAMC-B99EAD/2020\|EPI_ISL_673816\|20 | ···················· | ···················· | ···················· | ···················· | ···················· | ···················· |
| U | 3341 hCoV-19/England/CAMC-B99186/2020\|EPI_ISL_673817\|20 | ···················· | ···················· | ···················· | ···················· | ···················· | ···················· |
| U | 3342 hCoV-19/England/CAMC-B98907/2020\|EPI_ISL_673818\|20 | ···················· | ···················· | ···················· | ···················· | ···················· | ···················· |
| U | 3343 hCoV-19/England/CAMC-B9A64A/2020\|EPI_ISL_673819\|20 | ···················· | ···················· | ···················· | ···················· | ···················· | ···················· |
| U | 3344 hCoV-19/England/MILK-B94D4B/2020\|EPI_ISL_676018\|20 | ···················· | ···················· | ···················· | ···················· | ···················· | ···················· |
| U | 3345 hCoV-19/England/MILK-B965D8/2020\|EPI_ISL_676019\|20 | ···················· | ···················· | ···················· | ···················· | ···················· | ···················· |
| U | 3346 hCoV-19/England/MILK-B94ABD/2020\|EPI_ISL_676021\|20 | ···················· | ···················· | ···················· | ···················· | ···················· | ···················· |
| U | 3347 hCoV-19/England/MILK-B94D3C/2020\|EPI_ISL_676022\|20 | ···················· | ···················· | ···················· | ···················· | ···················· | ···················· |
| U | 3348 hCoV-19/England/MILK-B942C4/2020\|EPI_ISL_676023\|20 | ···················· | ···················· | ···················· | ···················· | ···················· | ···················· |
| U | 3349 hCoV-19/England/MILK-B94A08/2020\|EPI_ISL_676024\|20 | ···················· | ···················· | ···················· | ···················· | ···················· | ···················· |
| U | 3350 hCoV-19/England/MILK-B94491/2020\|EPI_ISL_676025\|20 | ···················· | ···················· | ···················· | ···················· | ···················· | ···················· |
| U | 3351 hCoV-19/England/MILK-B95DA4/2020\|EPI_ISL_676027\|20 | ···················· | ·A·················· | ···················· | ···················· | ···················· | ···················· |
| U | 3352 hCoV-19/England/MILK-B94E84/2020\|EPI_ISL_676028\|20 | ···················· | ···················· | ···················· | ···················· | ···················· | ···················· |
| U | 3353 hCoV-19/England/MILK-B94279/2020\|EPI_ISL_676029\|20 | ···················· | ···················· | ···················· | ···················· | ···················· | ···················· |
| U | 3354 hCoV-19/England/MILK-B96848/2020\|EPI_ISL_676031\|20 | ···················· | ···················· | ···················· | ···················· | ···················· | ···················· |
| U | 3355 hCoV-19/England/MILK-B9616B/2020\|EPI_ISL_676033\|20 | ···················· | ···················· | ···················· | ···················· | ···················· | ···················· |
| U | 3356 hCoV-19/England/MILK-B96A15/2020\|EPI_ISL_676035\|20 | ···················· | ···················· | ···················· | ···················· | ···················· | ···················· |
| U | 3357 hCoV-19/England/MILK-B96392/2020\|EPI_ISL_676037\|20 | ···················· | ·A·················· | ···················· | ···················· | ···················· | ···A················ |
| U | 3358 hCoV-19/England/MILK-B94D0F/2020\|EPI_ISL_676038\|20 | ···················· | ···················· | ···················· | ···················· | ···················· | ···················· |
| U | 3359 hCoV-19/England/MILK-B95DE0/2020\|EPI_ISL_676040\|20 | ···················· | ···················· | ···················· | ···················· | ···················· | ···················· |
| U | 3360 hCoV-19/England/MILK-B96A33/2020\|EPI_ISL_676041\|20 | ···················· | ·A·················· | ···················· | ···················· | ···················· | ···A················ |
| U | 3361 hCoV-19/England/MILK-B966D5/2020\|EPI_ISL_676042\|20 | ···················· | ···················· | ···················· | ···················· | ···················· | ···················· |
| U | 3362 hCoV-19/England/MILK-B96040/2020\|EPI_ISL_676043\|20 | ···················· | ···················· | ···················· | ···················· | ···················· | ···················· |
| U | 3363 hCoV-19/England/MILK-B94965/2020\|EPI_ISL_676044\|20 | ···················· | ···················· | ···················· | ···················· | ···················· | ···················· |
| U | 3364 hCoV-19/England/MILK-B96954/2020\|EPI_ISL_676046\|20 | ···················· | ···················· | ···················· | ···················· | ···················· | ···················· |
| U | 3365 hCoV-19/England/MILK-B96B12/2020\|EPI_ISL_676047\|20 | ···················· | ···················· | ···················· | ···················· | ···················· | ···················· |
| U | 3366 hCoV-19/England/MILK-B969BE/2020\|EPI_ISL_676048\|20 | ···················· | ···················· | ···················· | ···················· | ···················· | ···················· |
| U | 3367 hCoV-19/England/MILK-B94473/2020\|EPI_ISL_676049\|20 | ···················· | ···················· | ···················· | ···················· | ···················· | ···················· |
| U | 3368 hCoV-19/England/MILK-B95D4A/2020\|EPI_ISL_676050\|20 | ···················· | ···················· | ···················· | ···················· | ···················· | ···················· |
| U | 3369 hCoV-19/England/MILK-B96927/2020\|EPI_ISL_676051\|20 | ···················· | ···················· | ···················· | ···················· | ···················· | ···················· |
| U | 3370 hCoV-19/England/MILK-B968C0/2020\|EPI_ISL_676052\|20 | ···················· | ···················· | ···················· | ···················· | ···················· | ···················· |
| U | 3371 hCoV-19/England/MILK-B960B9/2020\|EPI_ISL_676053\|20 | ···················· | ·A·················· | ···················· | ···················· | ···················· | ···A················ |
| U | 3372 hCoV-19/England/MILK-B965BA/2020\|EPI_ISL_676056\|20 | ···················· | ···················· | ···················· | ···················· | ···················· | ···················· |
| U | 3373 hCoV-19/England/MILK-B95F62/2020\|EPI_ISL_676057\|20 | ···················· | ···················· | ···················· | ···················· | ···················· | ···················· |
| U | 3374 hCoV-19/England/MILK-B960F5/2020\|EPI_ISL_676058\|20 | ···················· | ···················· | ···················· | ···················· | ···················· | ···················· |
| U | 3375 hCoV-19/England/MILK-B93D00/2020\|EPI_ISL_676059\|20 | ···················· | ····A··············· | ···················· | ···················· | ···················· | ···················· |
| U | 3376 hCoV-19/England/MILK-B95DD1/2020\|EPI_ISL_676060\|20 | ···················· | ···················· | ···················· | ···················· | ···················· | ···················· |
| U | 3377 hCoV-19/England/MILK-B94D5A/2020\|EPI_ISL_676061\|20 | ···················· | ···················· | ···················· | ···················· | ···················· | ···················· |
| U | 3378 hCoV-19/England/MILK-B9473E/2020\|EPI_ISL_676062\|20 | ···················· | ···················· | ···················· | ···················· | ···················· | ···················· |
| U | 3379 hCoV-19/England/MILK-B942F1/2020\|EPI_ISL_676063\|20 | ···················· | ···················· | ···················· | ···················· | ···················· | ···················· |
| U | 3380 hCoV-19/England/MILK-B9431C/2020\|EPI_ISL_676067\|20 | ···················· | ···················· | ···················· | ···················· | ···················· | ···················· |
| U | 3381 hCoV-19/England/MILK-B94E66/2020\|EPI_ISL_676068\|20 | ···················· | ···················· | ···················· | ···················· | ···················· | ···················· |
| U | 3382 hCoV-19/England/MILK-B9680C/2020\|EPI_ISL_676069\|20 | ···················· | ···················· | ···················· | ···················· | ···················· | ···················· |
| U | 3383 hCoV-19/England/MILK-B966E4/2020\|EPI_ISL_676070\|20 | ···················· | ···················· | ···················· | ···················· | ···················· | ···················· |
| U | 3384 hCoV-19/England/MILK-B94A17/2020\|EPI_ISL_676071\|20 | ···················· | ···················· | ···················· | ···················· | ···················· | ···················· |
| U | 3385 hCoV-19/England/MILK-B942E2/2020\|EPI_ISL_676073\|20 | ···················· | ···················· | ···················· | ···················· | ···················· | ···················· |
| U | 3386 hCoV-19/England/MILK-B96198/2020\|EPI_ISL_676075\|20 | ···················· | ···················· | ···················· | ···················· | ···················· | ···················· |
| U | 3387 hCoV-19/England/MILK-B94B23/2020\|EPI_ISL_676076\|20 | ···················· | ···················· | ···················· | ···················· | ···················· | ···················· |
| U | 3388 hCoV-19/England/MILK-B948E0/2020\|EPI_ISL_676077\|20 | ···················· | ···················· | ···················· | ···················· | ···················· | ···················· |
| U | 3389 hCoV-19/England/MILK-B94CB7/2020\|EPI_ISL_676079\|20 | ···················· | ···················· | ···················· | ···················· | ···················· | ···················· |
| U | 3390 hCoV-19/England/MILK-B969FA/2020\|EPI_ISL_676081\|20 | ···················· | ···················· | ···················· | ···················· | ···················· | ···················· |
| U | 3391 hCoV-19/England/MILK-B94A9F/2020\|EPI_ISL_676082\|20 | ···················· | ···················· | ···················· | ···················· | ···················· | ···················· |
| U | 3392 hCoV-19/England/MILK-B95D77/2020\|EPI_ISL_676083\|20 | ···················· | ···················· | ···················· | ···················· | ······T············· | ···················· |
| U | 3393 hCoV-19/England/MILK-B95D0E/2020\|EPI_ISL_676085\|20 | ···················· | ···················· | ···················· | ···················· | ···················· | ···················· |
| U | 3394 hCoV-19/England/MILK-B95D3B/2020\|EPI_ISL_676086\|20 | ···················· | ···················· | ···················· | ···················· | ···················· | ···················· |
| U | 3395 hCoV-19/England/MILK-B9440A/2020\|EPI_ISL_676087\|20 | ···················· | ···················· | ···················· | ···················· | ···················· | ···················· |
| U | 3396 hCoV-19/England/MILK-B9423D/2020\|EPI_ISL_676088\|20 | ···················· | ···················· | ···················· | ···················· | ······T············· | ···················· |
| U | 3397 hCoV-19/England/MILK-B94C5D/2020\|EPI_ISL_676089\|20 | ···················· | ···················· | ···················· | ···················· | ···················· | ···················· |
| U | 3398 hCoV-19/England/MILK-B96B4F/2020\|EPI_ISL_676090\|20 | ···················· | ···················· | ···················· | ···················· | ······T············· | ···················· |

| | | | | | | | |
|---|---|---|---|---|---|---|---|
| U | 3399 hCoV-19/England/MILK-B9667B/2020\|EPI_ISL_676091\|20 | ···················· | ···················· | ···················· | ···················· | ···················· | ···················· |
| U | 3400 hCoV-19/England/MILK-B96B30/2020\|EPI_ISL_676092\|20 | ···················· | ······A············· | ···················· | ···················· | ···················· | ···················· |
| U | 3401 hCoV-19/England/MILK-B94789/2020\|EPI_ISL_676094\|20 | ···················· | ···················· | ···················· | ···················· | ···················· | ···················· |
| U | 3402 hCoV-19/England/MILK-B94ACC/2020\|EPI_ISL_676096\|20 | ···················· | ···················· | ···················· | ···················· | ···················· | ···················· |
| U | 3403 hCoV-19/England/MILK-B96945/2020\|EPI_ISL_676097\|20 | ···················· | ···················· | ···················· | ···················· | ···················· | ···················· |
| U | 3404 hCoV-19/England/MILK-B96A24/2020\|EPI_ISL_676099\|20 | ···················· | ···················· | ···················· | ···················· | ···················· | ···················· |
| U | 3405 hCoV-19/USA/NY-Wadsworth-37605-01/2020\|EPI_ISL_676 | ···················· | ···················· | ···················· | ···················· | ···················· | ···················· |
| U | 3406 hCoV-19/USA/NY-Wadsworth-38148-01/2020\|EPI_ISL_676 | ···················· | ···················· | ···················· | ···················· | ···················· | ···················· |
| U | 3407 hCoV-19/USA/NY-Wadsworth-38360-01/2020\|EPI_ISL_676 | ···················· | ···················· | ···················· | ···················· | ···················· | ···················· |
| U | 3408 hCoV-19/USA/NY-Wadsworth-38618-01/2020\|EPI_ISL_676 | ···················· | ···················· | ···················· | ···················· | ···················· | ···················· |
| U | 3409 hCoV-19/USA/NY-Wadsworth-38627-01/2020\|EPI_ISL_676 | ···················· | ···················· | ···················· | ···················· | ···················· | ···················· |
| U | 3410 hCoV-19/USA/NY-Wadsworth-38888-01/2020\|EPI_ISL_676 | ···················· | ···················· | ···················· | ···················· | ···················· | ···················· |
| U | 3411 hCoV-19/USA/NY-Wadsworth-38889-01/2020\|EPI_ISL_676 | ···················· | ···················· | ···················· | ···················· | ···················· | ···················· |
| U | 3412 hCoV-19/USA/NY-Wadsworth-39491-01/2020\|EPI_ISL_676 | ···················· | ···················· | ···················· | ···················· | ···················· | ···················· |
| U | 3413 hCoV-19/USA/NY-Wadsworth-39826-01/2020\|EPI_ISL_676 | ···················· | ···················· | ···················· | ···················· | ···················· | ···················· |
| U | 3414 hCoV-19/USA/CO-CDPHE-2008161334/2020\|EPI_ISL_67730 | ········T··········· | ···················· | ···················· | ···················· | ···················· | ···················· |
| U | 3415 hCoV-19/USA/CO-CDPHE-2008161388/2020\|EPI_ISL_67730 | ···················· | ···················· | ···················· | ···················· | ···················· | ···················· |
| U | 3416 hCoV-19/USA/CO-CDPHE-2008170014/2020\|EPI_ISL_67730 | ········T··········· | ···················· | ···················· | ···················· | ···················· | ···················· |
| U | 3417 hCoV-19/USA/CO-CDPHE-2008170021/2020\|EPI_ISL_67730 | ···················· | ···················· | ···················· | ···················· | ···················· | ···················· |
| U | 3418 hCoV-19/USA/CO-CDPHE-2008170075/2020\|EPI_ISL_67730 | ···················· | ···················· | ···················· | ···················· | ···················· | ···················· |
| U | 3420 hCoV-19/USA/CO-CDPHE-2008170179/2020\|EPI_ISL_67730 | ···················· | ·A·················· | ···················· | ···················· | ···················· | ···················· |
| U | 3422 hCoV-19/USA/CO-CDPHE-2008170936/2020\|EPI_ISL_67730 | ···················· | ···················· | ···················· | ···················· | ···················· | ···················· |
| U | 3423 hCoV-19/USA/CO-CDPHE-2008181589/2020\|EPI_ISL_67730 | ···················· | ···················· | ···················· | ···················· | ···················· | ···················· |
| U | 3424 hCoV-19/USA/CO-CDPHE-2009090318/2020\|EPI_ISL_67731 | ···················· | ···················· | ···················· | ···················· | ···················· | ···················· |
| U | 3426 hCoV-19/USA/WI-UW-1940/2020\|EPI_ISL_677349\|2020-10 | ···················· | ···················· | ···················· | ···················· | ···················· | ···················· |
| U | 3427 hCoV-19/USA/WI-UW-1941/2020\|EPI_ISL_677350\|2020-09 | ···················· | ···················· | ···················· | ···················· | ···················· | ···················· |
| U | 3428 hCoV-19/USA/WI-UW-1942/2020\|EPI_ISL_677351\|2020-10 | ···················· | ···················· | ···················· | ···················· | ···················· | ···················· |
| U | 3429 hCoV-19/USA/WI-UW-1944/2020\|EPI_ISL_677352\|2020-07 | ···················· | ·A·················· | ···················· | ···················· | ···················· | ···················· |
| U | 3430 hCoV-19/USA/IL-UW-1945/2020\|EPI_ISL_677353\|2020-09 | ···················· | ···················· | ···················· | ···················· | ···················· | ···················· |
| U | 3431 hCoV-19/USA/IL-UW-1946/2020\|EPI_ISL_677354\|2020-09 | ···················· | ···················· | ···················· | ···················· | ···················· | ···················· |
| U | 3432 hCoV-19/USA/WI-UW-1947/2020\|EPI_ISL_677355\|2020-09 | ···················· | ···················· | ···················· | ···················· | ···················· | ···················· |
| U | 3435 hCoV-19/USA/IL-UW-1950/2020\|EPI_ISL_677358\|2020-09 | ···················· | ···················· | ···················· | ···················· | ···················· | ···················· |
| U | 3437 hCoV-19/USA/WI-UW-1952/2020\|EPI_ISL_677360\|2020-09 | ···················· | ···················· | ···················· | ···················· | ···················· | ···················· |
| U | 3439 hCoV-19/USA/WI-UW-1954/2020\|EPI_ISL_677362\|2020-09 | ···················· | ···················· | ···················· | ···················· | ···················· | ···················· |
| U | 3440 hCoV-19/USA/IL-UW-1955/2020\|EPI_ISL_677363\|2020-09 | ···················· | ···················· | ···················· | ···················· | ···················· | ···················· |
| U | 3442 hCoV-19/USA/IL-UW-1957/2020\|EPI_ISL_677365\|2020-09 | ···················· | ···················· | ···················· | ···················· | ···················· | ···················· |
| U | 3443 hCoV-19/USA/WI-UW-1958/2020\|EPI_ISL_677366\|2020-09 | ···················· | ···················· | ···················· | ···················· | ···················· | ···················· |
| U | 3444 hCoV-19/USA/IL-UW-1959/2020\|EPI_ISL_677367\|2020-09 | ···················· | ·A·················· | ···················· | ···················· | ···················· | ···················· |
| U | 3447 hCoV-19/USA/WI-UW-1962/2020\|EPI_ISL_677370\|2020-10 | ···················· | ···················· | ···················· | ···················· | ···················· | ···················· |
| U | 3448 hCoV-19/USA/WI-UW-1963/2020\|EPI_ISL_677371\|2020-09 | ···················· | ···················· | ···················· | ···················· | ···················· | ···················· |
| U | 3449 hCoV-19/USA/WI-UW-1964/2020\|EPI_ISL_677372\|2020-09 | ···················· | ··C················· | ···················· | ···················· | ···················· | ···················· |
| U | 3451 hCoV-19/USA/WI-UW-1966/2020\|EPI_ISL_677374\|2020-09 | ···················· | ···················· | ···················· | ···················· | ···················· | ···················· |
| U | 3452 hCoV-19/USA/WI-UW-1967/2020\|EPI_ISL_677375\|2020-09 | ···················· | ···················· | ···················· | ···················· | ···················· | ···················· |
| U | 3453 hCoV-19/USA/WI-UW-1968/2020\|EPI_ISL_677376\|2020-09 | ···················· | ··N················· | ···················· | ···················· | ···················· | ···················· |
| U | 3465 hCoV-19/USA/IL-UW-1984/2020\|EPI_ISL_677392\|2020-09 | ···················· | ···················· | ···················· | ···················· | ···················· | ···················· |
| U | 3472 hCoV-19/South Africa/KRISP-K005307/2020\|EPI_ISL_67 | ···················· | ···················· | ···················· | ···················· | ···················· | ···················· |
| U | 3473 hCoV-19/South Africa/KRISP-K005308/2020\|EPI_ISL_67 | ···················· | ···················· | ···················· | ···················· | ···················· | ···················· |
| U | 3474 hCoV-19/South Africa/KRISP-K005309/2020\|EPI_ISL_67 | ···················· | ···················· | ···················· | ···················· | ···················· | ···················· |
| U | 3475 hCoV-19/South Africa/KRISP-K005310/2020\|EPI_ISL_67 | ···················· | ·A·················· | ···················· | ···················· | ···················· | ···················· |
| U | 3476 hCoV-19/South Africa/KRISP-K005312/2020\|EPI_ISL_67 | ···················· | ·A·················· | ···················· | ···················· | ···················· | ···················· |
| U | 3477 hCoV-19/South Africa/KRISP-K005313/2020\|EPI_ISL_67 | ···················· | ···················· | ···················· | ···················· | ···················· | ···················· |
| U | 3478 hCoV-19/South Africa/KRISP-K005314/2020\|EPI_ISL_67 | ···················· | ···················· | ···················· | ···················· | ···················· | ···················· |
| U | 3479 hCoV-19/South Africa/KRISP-K005316/2020\|EPI_ISL_67 | ···················· | ···················· | ···················· | ···················· | ···················· | ···················· |
| U | 3480 hCoV-19/South Africa/KRISP-K005317/2020\|EPI_ISL_67 | ···················· | ···················· | ···················· | ···················· | ···················· | ···················· |
| U | 3481 hCoV-19/South Africa/KRISP-K005318/2020\|EPI_ISL_67 | ···················· | ···················· | ···················· | ···················· | ···················· | ···················· |
| U | 3482 hCoV-19/South Africa/KRISP-K005320/2020\|EPI_ISL_67 | ···················· | ·A·················· | ···················· | ···················· | ···················· | ···················· |
| U | 3483 hCoV-19/South Africa/KRISP-K005322/2020\|EPI_ISL_67 | ···················· | ···················· | ···················· | ···················· | ···················· | ···················· |
| U | 3484 hCoV-19/South Africa/KRISP-K005323/2020\|EPI_ISL_67 | ···················· | ···················· | ···················· | ···················· | ···················· | ···················· |
| U | 3485 hCoV-19/South Africa/KRISP-K005324/2020\|EPI_ISL_67 | ···················· | ···················· | ···················· | ···················· | ···················· | ···················· |
| U | 3486 hCoV-19/South Africa/KRISP-K005326/2020\|EPI_ISL_67 | ···················· | ···················· | ···················· | ···················· | ···················· | ···················· |
| U | 3487 hCoV-19/South Africa/KRISP-K005328/2020\|EPI_ISL_67 | ···················· | ···················· | ···················· | ···················· | ···················· | ···················· |
| U | 3488 hCoV-19/South Africa/KRISP-K005331/2020\|EPI_ISL_67 | ···················· | ·A·················· | ···················· | ···················· | ···················· | ···················· |
| U | 3489 hCoV-19/South Africa/KRISP-K005332/2020\|EPI_ISL_67 | ···················· | ·A·················· | ···················· | ···················· | ···················· | ···················· |
| U | 3490 hCoV-19/South Africa/KRISP-K005334/2020\|EPI_ISL_67 | ···················· | ···················· | ···················· | ···················· | ···················· | ···················· |
| U | 3491 hCoV-19/South Africa/KRISP-K005335/2020\|EPI_ISL_67 | ···················· | ·A·················· | ···················· | ···················· | ···················· | ···················· |
| U | 3492 hCoV-19/South Africa/KRISP-K005336/2020\|EPI_ISL_67 | ···················· | ···················· | ···················· | ···················· | ···················· | ···················· |
| U | 3495 hCoV-19/South Africa/KRISP-EC-K005356/2020\|EPI_ISL | ···················· | ···················· | ···················· | ···················· | ···················· | ···················· |
| U | 3496 hCoV-19/England/204521764/2020\|EPI_ISL_678645\|2020 | ···················· | ···················· | ···················· | ···················· | ···················· | ···················· |
| U | 3497 hCoV-19/England/204561387/2020\|EPI_ISL_678685\|2020 | ···················· | ···················· | ···················· | ···················· | ···················· | ···A················ |
| U | 3498 hCoV-19/England/204561474/2020\|EPI_ISL_678687\|2020 | ···················· | ···················· | ···················· | ···················· | ···················· | ···R················ |
| U | 3499 hCoV-19/England/BIRM-6A787/2020\|EPI_ISL_679082\|202 | ···················· | ···················· | ···················· | ···················· | ······T············· | ···················· |
| U | 3500 hCoV-19/England/BIRM-6A7B4/2020\|EPI_ISL_679084\|202 | ···················· | ···················· | ···················· | ···················· | ···················· | ···················· |
| U | 3501 hCoV-19/England/BIRM-6A7C3/2020\|EPI_ISL_679085\|202 | ···················· | ···················· | ···················· | ···················· | ···················· | ···················· |
| U | 3502 hCoV-19/England/BIRM-6A7E1/2020\|EPI_ISL_679086\|202 | ···················· | ···················· | ···················· | ···················· | ···················· | ···················· |
| U | 3503 hCoV-19/England/BIRM-6A81B/2020\|EPI_ISL_679089\|202 | ···················· | ···················· | ···················· | ···················· | ···················· | ···················· |
| U | 3504 hCoV-19/England/NOTT-11707E/2020\|EPI_ISL_679924\|20 | ···················· | ···················· | ···················· | ···················· | ···················· | ···················· |
| U | 3505 hCoV-19/England/NOTT-117366/2020\|EPI_ISL_679925\|20 | ···················· | ···················· | ···················· | ···················· | ···················· | ···················· |
| U | 3506 hCoV-19/England/NOTT-1185D8/2020\|EPI_ISL_679926\|20 | ···················· | ···················· | ···················· | ···················· | ···················· | ···················· |
| U | 3507 hCoV-19/England/NOTT-119C2D/2020\|EPI_ISL_679928\|20 | ···················· | ···················· | ···················· | ···················· | ···················· | ···················· |
| U | 3508 hCoV-19/England/NOTT-119CB4/2020\|EPI_ISL_679929\|20 | ···················· | ···················· | ···················· | ···················· | ···················· | ···················· |

| | | | | | | | |
|---|---|---|---|---|---|---|---|
| U | 3509 hCoV-19/England/NOTT-119CC3/2020\|EPI_ISL_679930\|20 | ···················· | ···················· | ···················· | ···················· | ···················· | ···················· |
| U | 3510 hCoV-19/England/NOTT-11A1E1/2020\|EPI_ISL_679932\|20 | ···················· | ···················· | ···················· | ···················· | ···················· | ···················· |
| U | 3511 hCoV-19/England/NOTT-11CC57/2020\|EPI_ISL_679933\|20 | ···················· | ···················· | ···················· | ···················· | ···················· | ···················· |
| U | 3512 hCoV-19/England/NOTT-11DB2C/2020\|EPI_ISL_679935\|20 | ···················· | ···················· | ···················· | ···················· | ···················· | ···················· |
| U | 3513 hCoV-19/England/NOTT-11DBA4/2020\|EPI_ISL_679936\|20 | ···················· | ···················· | ···················· | ···················· | ···················· | ···················· |
| U | 3514 hCoV-19/England/NOTT-11DC65/2020\|EPI_ISL_679937\|20 | ···················· | ···················· | ···················· | ···················· | ···················· | ···················· |
| U | 3515 hCoV-19/England/NOTT-11DD71/2020\|EPI_ISL_679938\|20 | ···················· | ···················· | ···················· | ···················· | ···················· | ···················· |
| U | 3516 hCoV-19/England/NOTT-11DE6F/2020\|EPI_ISL_679939\|20 | ···················· | ···················· | ···················· | ···················· | ···················· | ···················· |
| U | 3517 hCoV-19/England/NOTT-11DE7E/2020\|EPI_ISL_679940\|20 | ···················· | ···················· | ···················· | ···················· | ···················· | ···················· |
| U | 3518 hCoV-19/England/NOTT-11DE8D/2020\|EPI_ISL_679941\|20 | ···················· | ···················· | ···················· | ···················· | ···················· | ···················· |
| U | 3519 hCoV-19/England/NOTT-11DE9C/2020\|EPI_ISL_679942\|20 | ···················· | ···················· | ···················· | ···················· | ···················· | ···················· |
| U | 3520 hCoV-19/England/NOTT-11E94F/2020\|EPI_ISL_679944\|20 | ···················· | ···················· | ···················· | ···················· | ···················· | ···················· |
| U | 3521 hCoV-19/England/NOTT-11EC64/2020\|EPI_ISL_679945\|20 | ···················· | ···················· | ···················· | ···················· | ···················· | ···················· |
| U | 3522 hCoV-19/England/NOTT-11F173/2020\|EPI_ISL_679946\|20 | ···················· | ···················· | ···················· | ···················· | ···················· | ···················· |
| U | 3523 hCoV-19/England/NOTT-11F216/2020\|EPI_ISL_679951\|20 | ···················· | ···················· | ···················· | ···················· | ···················· | ···················· |
| U | 3524 hCoV-19/England/NOTT-11F234/2020\|EPI_ISL_679952\|20 | ···················· | ···················· | ···················· | ···················· | ···················· | ···················· |
| U | 3525 hCoV-19/England/NOTT-11F252/2020\|EPI_ISL_679953\|20 | ···················· | ···················· | ···················· | ···················· | ···················· | ···················· |
| U | 3526 hCoV-19/England/NOTT-11F261/2020\|EPI_ISL_679954\|20 | ···················· | ···················· | ···················· | ···················· | ···················· | ···················· |
| U | 3527 hCoV-19/England/NOTT-11F2AD/2020\|EPI_ISL_679957\|20 | ···················· | ···················· | ···················· | ···················· | ···················· | ···················· |
| U | 3528 hCoV-19/England/NOTT-11F2CB/2020\|EPI_ISL_679958\|20 | ···················· | ···················· | ···················· | ···················· | ···················· | ···················· |
| U | 3529 hCoV-19/England/NOTT-11F2F8/2020\|EPI_ISL_679960\|20 | ···················· | ···················· | ···················· | ···················· | ···················· | ···················· |
| U | 3530 hCoV-19/England/NOTT-11F322/2020\|EPI_ISL_679962\|20 | ···················· | ···················· | ···················· | ···················· | ···················· | ···················· |
| U | 3531 hCoV-19/England/NOTT-11F36E/2020\|EPI_ISL_679965\|20 | ···················· | ···················· | ···················· | ···················· | ···················· | ···················· |
| U | 3532 hCoV-19/England/NOTT-11F3C8/2020\|EPI_ISL_679967\|20 | ···················· | ···················· | ···················· | ···················· | ···················· | ···················· |
| U | 3533 hCoV-19/England/NOTT-11F3E6/2020\|EPI_ISL_679968\|20 | ···················· | ···················· | ···················· | ···················· | ···················· | ···················· |
| U | 3534 hCoV-19/England/QEUH-B2C1F1/2020\|EPI_ISL_680070\|20 | ···················· | ···················· | ···················· | ···················· | ·····T·············· | ···················· |
| U | 3535 hCoV-19/England/QEUH-B2C2A3/2020\|EPI_ISL_680076\|20 | ···················· | ···················· | ···················· | ···················· | ···················· | ···················· |
| U | 3539 hCoV-19/Wales/PHWC-48E7D4/2020\|EPI_ISL_680913\|2020 | ···················· | ···················· | ···················· | ···················· | ···················· | ···················· |
| U | 3540 hCoV-19/Wales/PHWC-48E7E3/2020\|EPI_ISL_680914\|2020 | ···················· | ···················· | ···················· | ···················· | ···················· | ···················· |
| U | 3541 hCoV-19/Wales/PHWC-48E7F2/2020\|EPI_ISL_680915\|2020 | ···················· | ···················· | ···················· | ···················· | ···················· | ···················· |
| U | 3542 hCoV-19/Wales/PHWC-48E80E/2020\|EPI_ISL_680916\|2020 | ···················· | ···················· | ···················· | ···················· | ···················· | ···················· |
| U | 3543 hCoV-19/Wales/PHWC-48E859/2020\|EPI_ISL_680920\|2020 | ···················· | ···················· | ···················· | ···················· | ···················· | ···················· |
| U | 3544 hCoV-19/Wales/PHWC-48E868/2020\|EPI_ISL_680921\|2020 | ···················· | ···················· | ···················· | ···················· | ···················· | ···················· |
| U | 3545 hCoV-19/Wales/PHWC-48E886/2020\|EPI_ISL_680923\|2020 | ···················· | ···················· | ···················· | ···················· | ···················· | ···················· |
| U | 3546 hCoV-19/Wales/PHWC-48E895/2020\|EPI_ISL_680924\|2020 | ···················· | ···················· | ···················· | ···················· | ···················· | ···················· |
| U | 3549 hCoV-19/Wales/PHWC-48E8C2/2020\|EPI_ISL_680927\|2020 | ···················· | ···················· | ···················· | ···················· | ···················· | ···················· |
| U | 3551 hCoV-19/Wales/PHWC-48E8E0/2020\|EPI_ISL_680929\|2020 | ···················· | ···················· | ···················· | ···················· | ···················· | ···················· |
| U | 3553 hCoV-19/Wales/PHWC-48E90B/2020\|EPI_ISL_680931\|2020 | ···················· | ···················· | ···················· | ···················· | ···················· | ···················· |
| U | 3554 hCoV-19/Wales/PHWC-48E91A/2020\|EPI_ISL_680932\|2020 | ···················· | ···················· | ···················· | ···················· | ···················· | ···················· |
| U | 3556 hCoV-19/Wales/PHWC-48E947/2020\|EPI_ISL_680935\|2020 | ···················· | ···················· | ···················· | ···················· | ···················· | ···················· |
| U | 3557 hCoV-19/Wales/PHWC-48E956/2020\|EPI_ISL_680936\|2020 | ···················· | ···················· | ···················· | ···················· | ···················· | ···················· |
| U | 3559 hCoV-19/Wales/PHWC-48E992/2020\|EPI_ISL_680940\|2020 | ···················· | ···················· | ···················· | ···················· | ···················· | ···················· |
| U | 3564 hCoV-19/Wales/PHWC-48EA08/2020\|EPI_ISL_680947\|2020 | ···················· | ···················· | ···················· | ···················· | ···················· | ···················· |
| U | 3565 hCoV-19/Wales/PHWC-48EA17/2020\|EPI_ISL_680948\|2020 | ···················· | ···················· | ···················· | ···················· | ···················· | ···················· |
| U | 3566 hCoV-19/Wales/PHWC-48EA26/2020\|EPI_ISL_680949\|2020 | ···················· | ···················· | ···················· | ···················· | ···················· | ···················· |
| U | 3568 hCoV-19/Wales/PHWC-48EA44/2020\|EPI_ISL_680951\|2020 | ···················· | ···················· | ···················· | ···················· | ···················· | ···················· |
| U | 3569 hCoV-19/Wales/PHWC-48EA53/2020\|EPI_ISL_680952\|2020 | ···················· | ···················· | ···················· | ···················· | ···················· | ···················· |
| U | 3570 hCoV-19/Wales/PHWC-48EA62/2020\|EPI_ISL_680953\|2020 | ···················· | ···················· | ···················· | ···················· | ···················· | ···················· |
| U | 3572 hCoV-19/Wales/PHWC-48EA9F/2020\|EPI_ISL_680956\|2020 | ···················· | ···················· | ···················· | ···················· | ···················· | ···················· |
| U | 3573 hCoV-19/Wales/PHWC-48EAAE/2020\|EPI_ISL_680957\|2020 | ···················· | ···················· | ···················· | ···················· | ···················· | ···················· |
| U | 3574 hCoV-19/Wales/PHWC-48EABD/2020\|EPI_ISL_680958\|2020 | ···················· | ···················· | ···················· | ···················· | ···················· | ···················· |
| U | 3576 hCoV-19/Wales/PHWC-48ECC6/2020\|EPI_ISL_680990\|2020 | ···················· | ···················· | ···················· | ···················· | ···················· | ···················· |
| U | 3577 hCoV-19/Wales/PHWC-48ECE4/2020\|EPI_ISL_680992\|2020 | ···················· | ···················· | ···················· | ···················· | ···················· | ···················· |
| U | 3578 hCoV-19/Wales/PHWC-48ECF3/2020\|EPI_ISL_680993\|2020 | ···················· | ···················· | ···················· | ···················· | ···················· | ···················· |
| U | 3579 hCoV-19/Wales/PHWC-48ED0F/2020\|EPI_ISL_680994\|2020 | ···················· | ···················· | ···················· | ···················· | ···················· | ···················· |
| U | 3580 hCoV-19/Wales/PHWC-48ED1E/2020\|EPI_ISL_680995\|2020 | ···················· | ···················· | ···················· | ···················· | ···················· | ···················· |
| U | 3581 hCoV-19/Wales/PHWC-48ED2D/2020\|EPI_ISL_680996\|2020 | ···················· | ···················· | ···················· | ···················· | ···················· | ···················· |
| U | 3582 hCoV-19/Wales/PHWC-48ED3C/2020\|EPI_ISL_680997\|2020 | ···················· | ···················· | ···················· | ···················· | ···················· | ···················· |
| U | 3583 hCoV-19/Wales/PHWC-48ED4B/2020\|EPI_ISL_680998\|2020 | ···················· | ···················· | ···················· | ···················· | ···················· | ···················· |
| U | 3584 hCoV-19/Wales/PHWC-48ED5A/2020\|EPI_ISL_680999\|2020 | ···················· | ···················· | ···················· | ···················· | ···················· | ···················· |
| U | 3585 hCoV-19/England/ALDP-B84467/2020\|EPI_ISL_674207\|20 | ···················· | ·A·················· | ···················· | ···················· | ···················· | ··A················· |
| U | 3586 hCoV-19/England/MILK-B8C47E/2020\|EPI_ISL_674209\|20 | ···················· | ···················· | ···················· | ···················· | ···················· | ···················· |
| U | 3587 hCoV-19/England/MILK-B887B5/2020\|EPI_ISL_674214\|20 | ···················· | ·A·················· | ···················· | ···················· | ···················· | ··A················· |
| U | 3588 hCoV-19/England/MILK-B8C405/2020\|EPI_ISL_674216\|20 | ···················· | ···················· | ···················· | ···················· | ···················· | ···················· |
| U | 3589 hCoV-19/England/ALDP-B840EB/2020\|EPI_ISL_674217\|20 | ···················· | ···················· | ···················· | ···················· | ···················· | ···················· |
| U | 3590 hCoV-19/England/MILK-B8C344/2020\|EPI_ISL_674219\|20 | ···················· | ···················· | ···················· | ···················· | ···················· | ···················· |
| U | 3591 hCoV-19/England/MILK-B8C4AB/2020\|EPI_ISL_674220\|20 | ···················· | ···················· | ···················· | ···················· | ···················· | ···················· |
| U | 3592 hCoV-19/England/MILK-B8881C/2020\|EPI_ISL_674221\|20 | ···················· | ···················· | ···················· | ···················· | ···················· | ···················· |
| U | 3593 hCoV-19/England/ALDP-B85457/2020\|EPI_ISL_674222\|20 | ···················· | ···················· | ···················· | ···················· | ···················· | ···················· |
| U | 3594 hCoV-19/England/ALDP-B850F9/2020\|EPI_ISL_674223\|20 | ···················· | ···················· | ···················· | ···················· | ···················· | ···················· |
| U | 3595 hCoV-19/England/ALDP-B84AED/2020\|EPI_ISL_674224\|20 | ···················· | ···················· | ···················· | ···················· | ···················· | ···················· |
| U | 3596 hCoV-19/England/MILK-B8C432/2020\|EPI_ISL_674225\|20 | ···················· | ···················· | ···················· | ···················· | ···················· | ···················· |
| U | 3597 hCoV-19/England/ALDP-B83F58/2020\|EPI_ISL_674226\|20 | ···················· | ···················· | ···················· | ···················· | ···················· | ···················· |
| U | 3598 hCoV-19/England/ALDP-B84B08/2020\|EPI_ISL_674228\|20 | ···················· | ···················· | ···················· | ···················· | ···················· | ···················· |
| U | 3599 hCoV-19/England/ALDP-B83E2E/2020\|EPI_ISL_674229\|20 | ···················· | ···················· | ···················· | ···················· | ···················· | ···················· |
| U | 3600 hCoV-19/England/MILK-B8BF14/2020\|EPI_ISL_674230\|20 | ···················· | ···················· | ···················· | ···················· | ···················· | ···················· |
| U | 3601 hCoV-19/England/ALDP-B84A83/2020\|EPI_ISL_674231\|20 | ···················· | ···················· | ···················· | ···················· | ···················· | ···················· |
| U | 3602 hCoV-19/England/MILK-B88375/2020\|EPI_ISL_674232\|20 | ···················· | ···················· | ···················· | ···················· | ···················· | ···················· |
| U | 3603 hCoV-19/England/ALDP-B853D2/2020\|EPI_ISL_674234\|20 | ···················· | ···················· | ···················· | ···················· | ·····T·············· | ···················· |
| U | 3604 hCoV-19/England/ALDP-B858D3/2020\|EPI_ISL_675541\|20 | ···················· | ···················· | ···················· | ···················· | ···················· | ···················· |

| | | | | | | | | |
|---|---|---|---|---|---|---|---|---|
| U | 3605 hCoV-19/England/ALDP-B85B43/2020\|EPI_ISL_675545\|20 | ···················· | ···················· | ···················· | ···················· | ···················· | ···················· | |
| U | 3606 hCoV-19/England/ALDP-B92493/2020\|EPI_ISL_675546\|20 | ···················· | ···················· | ···················· | ···················· | ···················· | ···················· | |
| U | 3607 hCoV-19/England/MILK-B89DB2/2020\|EPI_ISL_675547\|20 | ···················· | ···················· | ···················· | ···················· | ·····T·············· | ···················· | |
| U | 3608 hCoV-19/England/ALDP-B85B52/2020\|EPI_ISL_675548\|20 | ···················· | ·A·················· | ···················· | ···················· | ···················· | ···················· | |
| U | 3609 hCoV-19/England/MILK-B89D2B/2020\|EPI_ISL_675550\|20 | ···················· | ···················· | ···················· | ···················· | ···················· | ···················· | |
| U | 3610 hCoV-19/England/ALDP-B82B64/2020\|EPI_ISL_675551\|20 | ···················· | ·A·················· | ···················· | ···················· | ···················· | ··A················· | |
| U | 3611 hCoV-19/England/ALDP-B8582E/2020\|EPI_ISL_675552\|20 | ···················· | ···················· | ···················· | ···················· | ···················· | ···················· | |
| U | 3612 hCoV-19/England/ALDP-B85AFB/2020\|EPI_ISL_675553\|20 | ···················· | ···················· | ···················· | ···················· | ···················· | ···················· | |
| U | 3613 hCoV-19/England/ALDP-B83B18/2020\|EPI_ISL_675554\|20 | ···················· | ···················· | ···················· | ···················· | ···················· | ···················· | |
| U | 3614 hCoV-19/England/MILK-B89DEF/2020\|EPI_ISL_675556\|20 | ···················· | ···················· | ···················· | ···················· | ···················· | ················A··· | |
| U | 3615 hCoV-19/England/ALDP-B82B73/2020\|EPI_ISL_675557\|20 | ···················· | ···················· | ···················· | ···················· | ···················· | ···················· | |
| U | 3616 hCoV-19/England/ALDP-B92A28/2020\|EPI_ISL_675559\|20 | ···················· | ···················· | ···················· | ···················· | ···················· | ···················· | |
| U | 3617 hCoV-19/England/ALDP-B83134/2020\|EPI_ISL_675561\|20 | ···················· | ···················· | ···················· | ···················· | ···················· | ···················· | |
| U | 3618 hCoV-19/England/ALDP-B8532D/2020\|EPI_ISL_675563\|20 | ···················· | ···················· | ···················· | ···················· | ···················· | ···················· | |
| U | 3619 hCoV-19/England/MILK-B89D3A/2020\|EPI_ISL_675564\|20 | ···················· | ···················· | ···················· | ···················· | ···················· | ···················· | |
| U | 3620 hCoV-19/England/MILK-B89EEC/2020\|EPI_ISL_675565\|20 | ···················· | ···················· | ···················· | ···················· | ···················· | ···················· | |
| U | 3621 hCoV-19/USA/NY-Wadsworth-40040-01/2020\|EPI_ISL_676 | ···················· | ···················· | ···················· | ···················· | ···················· | ···················· | |
| U | 3622 hCoV-19/USA/NY-Wadsworth-63708-01/2020\|EPI_ISL_676 | ···················· | ···················· | ···················· | ···················· | ···················· | ···················· | |
| U | 3623 hCoV-19/USA/NY-Wadsworth-64197-01/2020\|EPI_ISL_676 | ···················· | ···················· | ···················· | ···················· | ···················· | ···················· | |
| U | 3624 hCoV-19/USA/NY-Wadsworth-65138-01/2020\|EPI_ISL_676 | ···················· | ···················· | ···················· | ···················· | ···················· | ···················· | |
| U | 3625 hCoV-19/USA/NY-Wadsworth-65679-01/2020\|EPI_ISL_676 | ···················· | ···················· | ···················· | ···················· | ···················· | ···················· | |
| U | 3626 hCoV-19/USA/NY-Wadsworth-65680-01/2020\|EPI_ISL_676 | ···················· | ···················· | ···················· | ···················· | ···················· | ···················· | |
| U | 3627 hCoV-19/USA/NY-Wadsworth-65682-01/2020\|EPI_ISL_676 | ···················· | ···················· | ···················· | ···················· | ···················· | ···················· | |
| U | 3628 hCoV-19/USA/NY-Wadsworth-65683-01/2020\|EPI_ISL_676 | ···················· | ···················· | ···················· | ···················· | ···················· | ···················· | |
| U | 3629 hCoV-19/USA/NY-Wadsworth-65746-01/2020\|EPI_ISL_676 | ···················· | ···················· | ···················· | ···················· | ···················· | ···················· | |
| U | 3630 hCoV-19/USA/NY-Wadsworth-65860-01/2020\|EPI_ISL_676 | ···················· | ···················· | ···················· | ···················· | ···················· | ···················· | |
| U | 3631 hCoV-19/USA/NY-Wadsworth-68191-01/2020\|EPI_ISL_676 | ···················· | ···················· | ···················· | ···················· | ···················· | ···················· | |
| U | 3632 hCoV-19/USA/NY-Wadsworth-68281-01/2020\|EPI_ISL_676 | ···················· | ···················· | ···················· | ···················· | ···················· | ···················· | |
| U | 3633 hCoV-19/USA/NY-Wadsworth-68391-01/2020\|EPI_ISL_676 | ···················· | ···················· | ···················· | ···················· | ···················· | ···················· | |
| U | 3634 hCoV-19/USA/NY-Wadsworth-69720-01/2020\|EPI_ISL_676 | ···················· | ···················· | ···················· | ···················· | ···················· | ···················· | |
| U | 3635 hCoV-19/USA/NY-Wadsworth-70396-02/2020\|EPI_ISL_676 | ···················· | ···················· | ···················· | ···················· | ···················· | ···················· | |
| U | 3636 hCoV-19/USA/NY-Wadsworth-70464-01/2020\|EPI_ISL_676 | ···················· | ···················· | ···················· | ···················· | ···················· | ···················· | |
| U | 3637 hCoV-19/USA/NY-Wadsworth-71356-01/2020\|EPI_ISL_676 | ···················· | ···················A | ···················· | ···················· | ···················· | ···················· | 1 |
| U | 3638 hCoV-19/USA/NY-Wadsworth-72883-01/2020\|EPI_ISL_676 | ···················· | ···················· | ···················· | ···················· | ···················· | ···················· | |
| U | 3639 hCoV-19/USA/NY-Wadsworth-73244-01/2020\|EPI_ISL_676 | ···················· | ···················· | ···················· | ···················· | ···················· | ···················· | |
| U | 3640 hCoV-19/USA/NY-Wadsworth-073410-01/2020\|EPI_ISL_67 | ···················· | ···················· | ···················· | ···················· | ···················· | ···················· | |
| U | 3641 hCoV-19/USA/NY-Wadsworth-74236-01/2020\|EPI_ISL_676 | ···················· | ···················· | ···················· | ···················· | ···················· | ···················· | |
| U | 3642 hCoV-19/USA/NY-Wadsworth-75278-01/2020\|EPI_ISL_676 | ···················· | ···················· | ···················· | ···················· | ···················· | ···················· | |
| U | 3643 hCoV-19/USA/NY-Wadsworth-76300-01/2020\|EPI_ISL_676 | ···················· | ···················· | ···················· | ···················· | ···················· | ···················· | |
| U | 3644 hCoV-19/USA/NY-Wadsworth-76305-01/2020\|EPI_ISL_676 | ···················· | ···················· | ···················· | ···················· | ···················· | ···················· | |
| U | 3645 hCoV-19/USA/NY-Wadsworth-76306-01/2020\|EPI_ISL_676 | ···················· | ···················· | ···················· | ···················· | ···················· | ···················· | |
| U | 3646 hCoV-19/USA/NY-Wadsworth-76313-01/2020\|EPI_ISL_676 | ···················· | ···················· | ···················· | ···················· | ···················· | ···················· | |
| U | 3647 hCoV-19/England/BIRM-69FBD/2020\|EPI_ISL_679010\|202 | ···················· | ···················· | ···················· | ···················· | ···················· | ···················· | |
| U | 3648 hCoV-19/England/BIRM-6A013/2020\|EPI_ISL_679013\|202 | ···················· | ···················· | ···················· | ···················· | ···················· | ···················· | |
| U | 3649 hCoV-19/England/BIRM-6A022/2020\|EPI_ISL_679014\|202 | ···················· | ···················· | ···················· | ···················· | ···················· | ···················· | |
| U | 3650 hCoV-19/England/BIRM-6A06E/2020\|EPI_ISL_679016\|202 | ···················· | ···················· | ···················· | ···················· | ···················· | ···················· | |
| U | 3651 hCoV-19/England/BIRM-6A07D/2020\|EPI_ISL_679017\|202 | ···················· | ···················· | ···················· | ···················· | ···················· | ···················· | |
| U | 3652 hCoV-19/England/BIRM-6A08C/2020\|EPI_ISL_679018\|202 | ···················· | ···················· | ···················· | ···················· | ···················· | ···················· | |
| U | 3653 hCoV-19/England/BIRM-6A09B/2020\|EPI_ISL_679019\|202 | ···················· | ···················· | ···················· | ···················· | ···················· | ···················· | |
| U | 3654 hCoV-19/England/BIRM-6A0C8/2020\|EPI_ISL_679021\|202 | ···················· | ·A·················· | ···················· | ···················· | ···················· | ··A················· | |
| U | 3655 hCoV-19/England/BIRM-6A0E6/2020\|EPI_ISL_679022\|202 | ···················· | ···················· | ···················· | ···················· | ···················· | ···················· | |
| U | 3656 hCoV-19/England/BIRM-6A12F/2020\|EPI_ISL_679024\|202 | ···················· | ···················· | ···················· | ···················· | ···················· | ···················· | |
| U | 3657 hCoV-19/England/BIRM-6A15C/2020\|EPI_ISL_679025\|202 | ···················· | ···················· | ···················· | ···················· | ···················· | ···················· | |
| U | 3658 hCoV-19/England/BIRM-6A1A7/2020\|EPI_ISL_679026\|202 | ···················· | ·A·················· | ···················· | ···················· | ···················· | ··A················· | |
| U | 3659 hCoV-19/England/BIRM-6A1C5/2020\|EPI_ISL_679027\|202 | ···················· | ···················· | ···················· | ···················· | ···················· | ···················· | |
| U | 3660 hCoV-19/England/BIRM-6A1E3/2020\|EPI_ISL_679028\|202 | ···················· | ···················· | ···················· | ···················· | ···················· | ···················· | |
| U | 3661 hCoV-19/England/BIRM-6A23B/2020\|EPI_ISL_679030\|202 | ···················· | ···················· | ···················· | ···················· | ···················· | ···················· | |
| U | 3662 hCoV-19/England/BIRM-6A259/2020\|EPI_ISL_679032\|202 | ···················· | ···················· | ···················· | ···················· | ···················· | ···················· | |
| U | 3663 hCoV-19/England/BIRM-6A268/2020\|EPI_ISL_679033\|202 | ···················· | ···················· | ···················· | ···················· | ···················· | ···················· | |
| U | 3664 hCoV-19/England/BIRM-6A286/2020\|EPI_ISL_679034\|202 | ···················· | ···················· | ···················· | ···················· | ···················· | ···················· | |
| U | 3665 hCoV-19/England/BIRM-6A2A4/2020\|EPI_ISL_679036\|202 | ···················· | ·A·················· | ···················· | ···················· | ···················· | ··A················· | |
| U | 3666 hCoV-19/England/BIRM-6A2D1/2020\|EPI_ISL_679037\|202 | ···················· | ···················· | ···················· | ···················· | ···················· | ···················· | |
| U | 3667 hCoV-19/England/BIRM-6A2E0/2020\|EPI_ISL_679038\|202 | ···················· | ·A·················· | ···················· | ···················· | ···················· | ··A················· | |
| U | 3668 hCoV-19/England/BIRM-6A2FF/2020\|EPI_ISL_679039\|202 | ···················· | ···················· | ···················· | ···················· | ···················· | ···················· | |
| U | 3669 hCoV-19/England/BIRM-6A30B/2020\|EPI_ISL_679040\|202 | ···················· | ···················· | ···················· | ···················· | ···················· | ···················· | |
| U | 3670 hCoV-19/England/BIRM-6A31A/2020\|EPI_ISL_679041\|202 | ···················· | ···················· | ···················· | ···················· | ···················· | ···················· | |
| U | 3671 hCoV-19/England/BIRM-6A329/2020\|EPI_ISL_679042\|202 | ···················· | ···················· | ···················· | ···················· | ···················· | ···················· | |
| U | 3672 hCoV-19/England/BIRM-6A338/2020\|EPI_ISL_679043\|202 | ···················· | ···················· | ···················· | ···················· | ···················· | ···················· | |
| U | 3673 hCoV-19/England/BIRM-6A347/2020\|EPI_ISL_679044\|202 | ···················· | ···················· | ···················· | ···················· | ···················· | ···················· | |
| U | 3674 hCoV-19/England/BIRM-6A365/2020\|EPI_ISL_679045\|202 | ···················· | ···················· | ···················· | ···················· | ···················· | ···················· | |
| U | 3675 hCoV-19/England/BIRM-6A383/2020\|EPI_ISL_679047\|202 | ···················· | ···················· | ···················· | ···················· | ···················· | ···················· | |
| U | 3676 hCoV-19/England/BIRM-6A392/2020\|EPI_ISL_679048\|202 | ···················· | ···················· | ···················· | ···················· | ···················· | ···················· | |
| U | 3677 hCoV-19/England/BIRM-6A3A1/2020\|EPI_ISL_679049\|202 | ···················· | ···················· | ···················· | ···················· | ···················· | ···················· | |
| U | 3678 hCoV-19/England/BIRM-6A3B0/2020\|EPI_ISL_679050\|202 | ···················· | ···················· | ···················· | ···················· | ···················· | ···················· | |
| U | 3679 hCoV-19/England/BIRM-6A408/2020\|EPI_ISL_679053\|202 | ···················· | ·A·················· | ···················· | ···················· | ···················· | ··A················· | |
| U | 3680 hCoV-19/England/BIRM-6A417/2020\|EPI_ISL_679054\|202 | ···················· | ···················· | ···················· | ···················· | ···················· | ···················· | |
| U | 3681 hCoV-19/England/BIRM-6A426/2020\|EPI_ISL_679055\|202 | ···················· | ···················· | ···················· | ···················· | ···················· | ···················· | |
| U | 3682 hCoV-19/England/BIRM-6A444/2020\|EPI_ISL_679057\|202 | ···················· | ···················· | ···················· | ···················· | ···················· | ···················· | |
| U | 3683 hCoV-19/England/BIRM-6A453/2020\|EPI_ISL_679058\|202 | ···················· | ···················· | ···················· | ···················· | ···················· | ···················· | |
| U | 3684 hCoV-19/England/BIRM-6A462/2020\|EPI_ISL_679059\|202 | ···················· | ···················· | ···················· | ···················· | ···················· | ···················· | |

| | | | | | | | | |
|---|---|---|---|---|---|---|---|---|
| U | 3685 | hCoV-19/England/BIRM-6A471/2020\|EPI_ISL_679060\|202 | ···················· | ···················· | ···················· | ···················· | ···················· | ···················· |
| U | 3686 | hCoV-19/England/BIRM-6A480/2020\|EPI_ISL_679061\|202 | ···················· | ···················· | ···················· | ···················· | ···················· | ···················· |
| U | 3687 | hCoV-19/England/BIRM-6A49F/2020\|EPI_ISL_679062\|202 | ···················· | ···················· | ···················· | ···················· | ···················· | ···················· |
| U | 3688 | hCoV-19/England/BIRM-6A4BD/2020\|EPI_ISL_679063\|202 | ···················· | ···················· | ···················· | ···················· | ···················· | ···················· |
| U | 3689 | hCoV-19/England/BIRM-6A4EA/2020\|EPI_ISL_679065\|202 | ···················· | ···················· | ···················· | ···················· | ···················· | ···················· |
| U | 3690 | hCoV-19/England/BIRM-6A4F9/2020\|EPI_ISL_679066\|202 | ···················· | ···················· | ···················· | ···················· | ···················· | ···················· |
| U | 3691 | hCoV-19/England/BIRM-6A505/2020\|EPI_ISL_679067\|202 | ···················· | ···················· | ···················· | ···················· | ···················· | ···················· |
| U | 3692 | hCoV-19/England/BIRM-6A523/2020\|EPI_ISL_679069\|202 | ···················· | ···················· | ···················· | ···················· | ···················· | ···················· |
| U | 3693 | hCoV-19/England/BIRM-6A6E4/2020\|EPI_ISL_679072\|202 | ···················· | ···················· | ···················· | ···················· | ···················· | ···················· |
| U | 3694 | hCoV-19/England/BIRM-6A6F3/2020\|EPI_ISL_679073\|202 | ···················· | ···················· | ···················· | ···················· | ···················· | ···················· |
| U | 3695 | hCoV-19/England/BIRM-6A70F/2020\|EPI_ISL_679074\|202 | ···················· | ···················· | ···················· | ···················· | ···················· | ···················· |
| U | 3696 | hCoV-19/England/BIRM-6A71E/2020\|EPI_ISL_679075\|202 | ···················· | ···················· | ···················· | ···················· | ···················· | ···················· |
| U | 3697 | hCoV-19/England/BIRM-6A72D/2020\|EPI_ISL_679076\|202 | ···················· | ···················· | ···················· | ···················· | ···················· | ···················· |
| U | 3698 | hCoV-19/England/BIRM-6A73C/2020\|EPI_ISL_679077\|202 | ···················· | ···················· | ···················· | ···················· | ···················· | ···················· |
| U | 3699 | hCoV-19/England/BIRM-6A74B/2020\|EPI_ISL_679078\|202 | ···················· | ···················· | ···················· | ···················· | ···················· | ···················· |
| U | 3700 | hCoV-19/England/BIRM-6A75A/2020\|EPI_ISL_679079\|202 | ···················· | ···················· | ···················· | ···················· | ···················· | ···················· |
| U | 3701 | hCoV-19/England/BIRM-6A769/2020\|EPI_ISL_679080\|202 | ···················· | ···················· | ···················· | ···················· | ···················· | ···················· |
| U | 3702 | hCoV-19/England/BIRM-6A778/2020\|EPI_ISL_679081\|202 | ···················· | ···················· | ···················· | ···················· | ···················· | ···················· |
| U | 3703 | hCoV-19/England/QEUH-A018B2/2020\|EPI_ISL_680000\|20 | ···················· | ···················· | ···················· | ···················· | ···················· | ···················· |
| U | 3704 | hCoV-19/England/QEUH-A01AAD/2020\|EPI_ISL_680004\|20 | ···················· | ·A·················· | ···················· | ···················· | ···················· | ···················· |
| U | 3705 | hCoV-19/England/QEUH-A01B04/2020\|EPI_ISL_680006\|20 | ···················· | ···················· | ···················· | ···················· | ···················· | ···················· |
| U | 3706 | hCoV-19/England/QEUH-A01B22/2020\|EPI_ISL_680007\|20 | ···················· | ···················· | ···················· | ···················· | ···················· | ···················· |
| U | 3707 | hCoV-19/England/QEUH-A01B6E/2020\|EPI_ISL_680009\|20 | ···················· | ···················· | ···················· | ···················· | ···················· | ···················· |
| U | 3708 | hCoV-19/England/QEUH-A01BAA/2020\|EPI_ISL_680012\|20 | ·········T·········· | ···················· | ···················· | ···················· | ···················· | ···················· |
| U | 3709 | hCoV-19/England/QEUH-A9D6B1/2020\|EPI_ISL_680019\|20 | ···················· | ···················· | ···················· | ···················· | ···················· | ···················· |
| U | 3710 | hCoV-19/England/QEUH-A9D6EE/2020\|EPI_ISL_680020\|20 | ···················· | ···················· | ···················· | ···················· | ···················· | ···················· |
| U | 3711 | hCoV-19/England/QEUH-A9D921/2020\|EPI_ISL_680029\|20 | ···················· | ···················· | ···················· | ···················· | ···················· | ···················· |
| U | 3712 | hCoV-19/England/QEUH-B2BF43/2020\|EPI_ISL_680039\|20 | ···················· | ···················· | ···················· | ···················· | ···················· | ···················· |
| U | 3713 | hCoV-19/England/QEUH-B2BF70/2020\|EPI_ISL_680041\|20 | ···················· | ···················· | ···················· | ···················· | ···················· | ···················· |
| U | 3714 | hCoV-19/England/QEUH-B2BF9E/2020\|EPI_ISL_680043\|20 | ···················· | ···················· | ···················· | ···················· | ···················· | ···················· |
| U | 3715 | hCoV-19/England/QEUH-B2C012/2020\|EPI_ISL_680047\|20 | ···················· | ···················· | ···················· | ···················· | ···················· | ···················· |
| U | 3716 | hCoV-19/England/QEUH-B2C05E/2020\|EPI_ISL_680048\|20 | ···················· | ···················· | ···················· | ···················· | ···················· | ···················· |
| U | 3717 | hCoV-19/England/QEUH-B2C06D/2020\|EPI_ISL_680049\|20 | ···················· | ···················· | ···················· | ···················· | ···················· | ···················· |
| U | 3718 | hCoV-19/England/QEUH-B2C08B/2020\|EPI_ISL_680051\|20 | ···················· | ···················· | ···················· | ···················· | ···················· | ···················· |
| U | 3719 | hCoV-19/England/QEUH-B2C0A9/2020\|EPI_ISL_680053\|20 | ···················· | ···················· | ···················· | ···················· | ···················· | ···················· |
| U | 3720 | hCoV-19/England/QEUH-B2C0B8/2020\|EPI_ISL_680054\|20 | ···················· | ···················· | ···················· | ···················· | ···················· | ···················· |
| U | 3721 | hCoV-19/England/QEUH-B2C0D6/2020\|EPI_ISL_680056\|20 | ···················· | ···················· | ···················· | ···················· | ···················· | ···················· |
| U | 3722 | hCoV-19/England/QEUH-B2C0F4/2020\|EPI_ISL_680058\|20 | ···················· | ···················· | ···················· | ···················· | ···················· | ···················· |
| U | 3723 | hCoV-19/England/QEUH-B2C12E/2020\|EPI_ISL_680060\|20 | ···················· | ···················· | ···················· | ···················· | ···················· | ···················· |
| U | 3724 | hCoV-19/England/QEUH-B2C188/2020\|EPI_ISL_680064\|20 | ···················· | ···················· | ···················· | ···················· | ···················· | ···················· |
| U | 3725 | hCoV-19/England/QEUH-B2C1A6/2020\|EPI_ISL_680066\|20 | ···················· | ···················· | ···················· | ···················· | ···················· | ···················· |
| U | 3726 | hCoV-19/England/QEUH-B2C1B5/2020\|EPI_ISL_680067\|20 | ···················· | ···················· | ···················· | ···················· | ···················· | ···················· |
| U | 3728 | hCoV-19/USA/IL-UW-1621/2020\|EPI_ISL_681321\|2020-09 | ···················· | ···················· | ···················· | ···················· | ···················· | ···················· |
| U | 3729 | hCoV-19/USA/FL-UF-37/2020\|EPI_ISL_681322\|2020-06-3 | ···················· | ···················· | ···················· | ···················· | ···················· | ···················· |
| U | 3730 | hCoV-19/England/204881268/2020\|EPI_ISL_681324\|2020 | ···················· | ···················· | ···················· | ···················· | ······T············· | ···················· |
| U | 3731 | hCoV-19/England/MILK-BAB989/2020\|EPI_ISL_681325\|20 | ···················· | ···················· | ···················· | ···················· | ···················· | ···················· |
| U | 3732 | hCoV-19/England/MILK-BAA54A/2020\|EPI_ISL_681326\|20 | ···················· | ···················· | ···················· | ···················· | ···················· | ···················· |
| U | 3733 | hCoV-19/England/MILK-BAA9C6/2020\|EPI_ISL_681327\|20 | ···················· | ···················· | ···················· | ···················· | ···················· | ···················· |
| U | 3734 | hCoV-19/England/MILK-BABC26/2020\|EPI_ISL_681328\|20 | ···················· | ···················· | ···················· | ···················· | ···················· | ···················· |
| U | 3735 | hCoV-19/England/MILK-BAB1CD/2020\|EPI_ISL_681329\|20 | ···················· | ···················· | ···················· | ···················· | ···················· | ···················· |
| U | 3736 | hCoV-19/England/MILK-BAA1A0/2020\|EPI_ISL_681330\|20 | ···················· | ···················· | ···················· | ···················· | ···················· | ···················· |
| U | 3737 | hCoV-19/England/MILK-BA9E40/2020\|EPI_ISL_681331\|20 | ···················· | ···················· | ···················· | ···················· | ···················· | ···················· |
| U | 3738 | hCoV-19/England/MILK-BABE02/2020\|EPI_ISL_681332\|20 | ···················· | ···················· | ···················· | ···················· | ···················· | ···················· |
| U | 3739 | hCoV-19/England/MILK-BAC43C/2020\|EPI_ISL_681333\|20 | ···················· | ···················· | ···················· | ···················· | ······T············· | ···················· |
| U | 3740 | hCoV-19/England/MILK-BA9F6B/2020\|EPI_ISL_681334\|20 | ···················· | ···················· | ···················· | ···················· | ···················· | ···················· |
| U | 3741 | hCoV-19/England/MILK-BABBCF/2020\|EPI_ISL_681335\|20 | ···················· | ···················· | ···················· | ···················· | ···················· | ···················· |
| U | 3742 | hCoV-19/England/MILK-BAB97A/2020\|EPI_ISL_681336\|20 | ···················· | ···················· | ···················· | ···················· | ···················· | ···················· |
| U | 3743 | hCoV-19/England/MILK-BAA5D1/2020\|EPI_ISL_681337\|20 | ···················· | ···················· | ···················· | ···················· | ···················· | ···················· |
| U | 3744 | hCoV-19/England/MILK-BAA5E0/2020\|EPI_ISL_681338\|20 | ···················· | ···················· | ···················· | ···················· | ···················· | ···················· |
| U | 3745 | hCoV-19/England/MILK-BA9B76/2020\|EPI_ISL_681341\|20 | ···················· | ···················· | ···················· | ···················· | ···················· | ···················· |
| U | 3746 | hCoV-19/England/MILK-BAC45A/2020\|EPI_ISL_681342\|20 | ···················· | ···················· | ···················· | ···················· | ···················· | ···················· |
| U | 3747 | hCoV-19/England/MILK-BAA814/2020\|EPI_ISL_681343\|20 | ···················· | ···················· | ···················· | ···················· | ···················· | ···················· |
| U | 3748 | hCoV-19/England/MILK-BAB084/2020\|EPI_ISL_681344\|20 | ···················· | ···················· | ···················· | ···················· | ···················· | ···················· |
| U | 3749 | hCoV-19/England/MILK-BAAB93/2020\|EPI_ISL_681346\|20 | ···················· | ···················· | ···················· | ···················· | ······T············· | ···················· |
| U | 3750 | hCoV-19/England/MILK-BAB5EF/2020\|EPI_ISL_681348\|20 | ···················· | ···················· | ···················· | ···················· | ···················· | ···················· |
| U | 3751 | hCoV-19/England/MILK-BAAFF1/2020\|EPI_ISL_681349\|20 | ···················· | ···················· | ···················· | ···················· | ···················· | ···················· |
| U | 3752 | hCoV-19/England/MILK-BAA96C/2020\|EPI_ISL_681350\|20 | ···················· | ···················· | ···················· | ···················· | ···················· | ···················· |
| U | 3753 | hCoV-19/England/MILK-BAA3B9/2020\|EPI_ISL_681351\|20 | ···················· | ···················· | ···················· | ···················· | ···················· | ···················· |
| U | 3754 | hCoV-19/England/MILK-BAA3F5/2020\|EPI_ISL_681352\|20 | ···················· | ···················· | ···················· | ···················· | ···················· | ···················· |
| U | 3755 | hCoV-19/England/MILK-BABC53/2020\|EPI_ISL_681353\|20 | ···················· | ···················· | ···················· | ···················· | ···················· | ···················· |
| U | 3756 | hCoV-19/England/MILK-BA9EF5/2020\|EPI_ISL_681354\|20 | ···················· | ···················· | ···················· | ···················· | ···················· | ···················· |
| U | 3757 | hCoV-19/England/MILK-BA9B3A/2020\|EPI_ISL_681355\|20 | ···················· | ···················· | ···················· | ···················· | ···················· | ···················· |
| U | 3758 | hCoV-19/England/MILK-BAB44C/2020\|EPI_ISL_681356\|20 | ···················· | ···················· | ···················· | ···················· | ···················· | ···················· |
| U | 3759 | hCoV-19/England/MILK-BAA577/2020\|EPI_ISL_681357\|20 | ···················· | ···················· | ···················· | ···················· | ···················· | ···················· |
| U | 3760 | hCoV-19/England/MILK-BAB53A/2020\|EPI_ISL_681358\|20 | ···················· | ···················· | ···················· | ···················· | ···················· | ···················· |
| U | 3761 | hCoV-19/England/MILK-BACDD7/2020\|EPI_ISL_681359\|20 | ···················· | ···················· | ···················· | ···················· | ···················· | ···················· |
| U | 3762 | hCoV-19/England/MILK-BA9E5F/2020\|EPI_ISL_681360\|20 | ···················· | ···················· | ···················· | ···················· | ···················· | ···················· |
| U | 3763 | hCoV-19/England/MILK-BAA595/2020\|EPI_ISL_681361\|20 | ···················· | ···················· | ···················· | ···················· | ···················· | ···················· |
| U | 3764 | hCoV-19/England/MILK-BABBED/2020\|EPI_ISL_681362\|20 | ···················· | ···················· | ···················· | ···················· | ···················· | ···················· |
| U | 3765 | hCoV-19/England/MILK-BAB1DC/2020\|EPI_ISL_681363\|20 | ···················· | ···················· | ···················· | ···················· | ···················· | ···················· |

| | | | | | | | |
|---|---|---|---|---|---|---|---|
| U | 3766 hCoV-19/England/MILK-BABC08/2020\|EPI_ISL_681364\|20 | ···················· | ···················· | ···················· | ···················· | ···················· | ···················· |
| U | 3767 hCoV-19/England/MILK-BAA5B3/2020\|EPI_ISL_681365\|20 | ···················· | ···················· | ···················· | ···················· | ···················· | ···················· |
| U | 3768 hCoV-19/England/MILK-BAA216/2020\|EPI_ISL_681366\|20 | ···················· | ···················· | ···················· | ···················· | ···················· | ···················· |
| U | 3769 hCoV-19/England/MILK-BAB28E/2020\|EPI_ISL_681369\|20 | ···················· | ···················· | ···················· | ···················· | ···················· | ···················· |
| U | 3770 hCoV-19/England/MILK-BAB00C/2020\|EPI_ISL_681370\|20 | ···················· | ···················· | ···················· | ···················· | ···················· | ···················· |
| U | 3771 hCoV-19/England/MILK-BAB4C4/2020\|EPI_ISL_681371\|20 | ···················· | ···················· | ···················· | ···················· | ···················· | ···················· |
| U | 3772 hCoV-19/England/MILK-BAB2BB/2020\|EPI_ISL_681372\|20 | ···················· | ···················· | ···················· | ···················· | ···················· | ···················· |
| U | 3773 hCoV-19/England/MILK-BA9A3D/2020\|EPI_ISL_681373\|20 | ···················· | ···················· | ···················· | ···················· | ···················· | ···················· |
| U | 3774 hCoV-19/England/MILK-BA9F98/2020\|EPI_ISL_681374\|20 | ···················· | ···················· | ···················· | ···················· | ···················· | ···················· |
| U | 3775 hCoV-19/England/MILK-BAB95C/2020\|EPI_ISL_681375\|20 | ···················· | ···················· | ···················· | ···················· | ···················· | ···················· |
| U | 3776 hCoV-19/England/MILK-BAB9C5/2020\|EPI_ISL_681376\|20 | ···················· | ···················· | ···················· | ···················· | ···················· | ···················· |
| U | 3777 hCoV-19/England/MILK-BAB251/2020\|EPI_ISL_681378\|20 | ···················· | ···················· | ···················· | ···················· | ·····T·············· | ···················· |
| U | 3778 hCoV-19/England/MILK-BAB7BC/2020\|EPI_ISL_681379\|20 | ···················· | ···················· | ···················· | ···················· | ···················· | ···················· |
| U | 3779 hCoV-19/England/MILK-BA9E31/2020\|EPI_ISL_681380\|20 | ···················· | ···················· | ···················· | ···················· | ···················· | ···················· |
| U | 3780 hCoV-19/England/MILK-BAA88D/2020\|EPI_ISL_681382\|20 | ···················· | ···················· | ···················· | ···················· | ···················· | ···················· |
| U | 3781 hCoV-19/England/MILK-BAC627/2020\|EPI_ISL_681383\|20 | ···················· | ···················· | ···················· | ···················· | ···················· | ···················· |
| U | 3782 hCoV-19/England/MILK-BAA322/2020\|EPI_ISL_681384\|20 | ···················· | ···················· | ···················· | ···················· | ···················· | ···················· |
| U | 3783 hCoV-19/England/MILK-BABC17/2020\|EPI_ISL_681386\|20 | ···················· | ···················· | ···················· | ···················· | ···················· | ···················· |
| U | 3784 hCoV-19/England/MILK-BAAAA5/2020\|EPI_ISL_681387\|20 | ···················· | ···················· | ···················· | ···················· | ···················· | ···················· |
| U | 3785 hCoV-19/England/MILK-BAA1EC/2020\|EPI_ISL_681389\|20 | ···················· | ···················· | ···················· | ···················· | ···················· | ···················· |
| U | 3786 hCoV-19/England/MILK-BA99D6/2020\|EPI_ISL_681391\|20 | ···················· | ···················· | ···················· | ···················· | ···················· | ···················· |
| U | 3787 hCoV-19/England/MILK-BAAC09/2020\|EPI_ISL_681392\|20 | ···················· | ···················· | ···················· | ···················· | ···················· | ···················· |
| U | 3788 hCoV-19/England/MILK-BABAD1/2020\|EPI_ISL_681393\|20 | ···················· | ···················· | ···················· | ···················· | ···················· | ···················· |
| U | 3789 hCoV-19/England/MILK-BAAB48/2020\|EPI_ISL_681394\|20 | ···················· | ···················· | ···················· | ···················· | ···················· | ···················· |
| U | 3790 hCoV-19/England/MILK-BABE4E/2020\|EPI_ISL_681395\|20 | ···················· | ···················· | ···················· | ···················· | ···················· | ···················· |
| U | 3791 hCoV-19/England/MILK-BAB558/2020\|EPI_ISL_681396\|20 | ···················· | ···················· | ···················· | ···················· | ···················· | ···················· |
| U | 3792 hCoV-19/England/MILK-BAAC18/2020\|EPI_ISL_681397\|20 | ···················· | ···················· | ···················· | ···················· | ···················· | ···················· |
| U | 3793 hCoV-19/England/MILK-BAC690/2020\|EPI_ISL_681398\|20 | ···················· | ···················· | ···················· | ···················· | ···················· | ···················· |
| U | 3794 hCoV-19/England/MILK-BAA586/2020\|EPI_ISL_681399\|20 | ···················· | ···················· | ···················· | ···················· | ···················· | ···················· |
| U | 3795 hCoV-19/Denmark/DCGC-12470/2020\|EPI_ISL_682651\|202 | ···················· | ···················· | ···················· | ···················· | ···················· | ···················· |
| U | 3796 hCoV-19/Denmark/DCGC-12473/2020\|EPI_ISL_682654\|202 | ···················· | ···················· | ···················· | ···················· | ···················· | ···················· |
| U | 3797 hCoV-19/Denmark/DCGC-12474/2020\|EPI_ISL_682655\|202 | ···················· | ···················· | ···················· | ···················· | ···················· | ···················· |
| U | 3798 hCoV-19/Denmark/DCGC-12475/2020\|EPI_ISL_682656\|202 | ···················· | ···················· | ···················· | ···················· | ···················· | ···················· |
| U | 3799 hCoV-19/Denmark/DCGC-12476/2020\|EPI_ISL_682657\|202 | ···················· | ···················· | ···················· | ···················· | ···················· | ···················· |
| U | 3800 hCoV-19/Denmark/DCGC-12480/2020\|EPI_ISL_682661\|202 | ···················· | ···················· | ···················· | ···················· | ···················· | ···················· |
| U | 3801 hCoV-19/Denmark/DCGC-12481/2020\|EPI_ISL_682662\|202 | ···················· | ···················· | ···················· | ···················· | ···················· | ···················· |
| U | 3802 hCoV-19/Denmark/DCGC-12482/2020\|EPI_ISL_682663\|202 | ···················· | ···················· | ···················· | ···················· | ···················· | ···················· |
| U | 3803 hCoV-19/Denmark/DCGC-12483/2020\|EPI_ISL_682664\|202 | ···················· | ···················· | ···················· | ···················· | ···················· | ···················· |
| U | 3804 hCoV-19/Denmark/DCGC-12485/2020\|EPI_ISL_682666\|202 | ···················· | ···················· | ···················· | ···················· | ···················· | ···················· |
| U | 3805 hCoV-19/Denmark/DCGC-12486/2020\|EPI_ISL_682667\|202 | ···················· | ···················· | ···················· | ···················· | ···················· | ···················· |
| U | 3806 hCoV-19/Denmark/DCGC-12489/2020\|EPI_ISL_682670\|202 | ···················· | ···················· | ···················· | ···················· | ···················· | ···················· |
| U | 3807 hCoV-19/Denmark/DCGC-12491/2020\|EPI_ISL_682672\|202 | ···················· | ···················· | ···················· | ···················· | ···················· | ···················· |
| U | 3808 hCoV-19/Denmark/DCGC-12492/2020\|EPI_ISL_682673\|202 | ···················· | ···················· | ···················· | ···················· | ···················· | ···················· |
| U | 3809 hCoV-19/Denmark/DCGC-12493/2020\|EPI_ISL_682674\|202 | ···················· | ···················· | ···················· | ···················· | ···················· | ···················· |
| U | 3810 hCoV-19/Denmark/DCGC-12498/2020\|EPI_ISL_682679\|202 | ···················· | ···················· | ···················· | ···················· | ···················· | ···················· |
| U | 3811 hCoV-19/Denmark/DCGC-12499/2020\|EPI_ISL_682680\|202 | ···················· | ···················· | ···················· | ···················· | ···················· | ···················· |
| U | 3812 hCoV-19/Denmark/DCGC-12500/2020\|EPI_ISL_682681\|202 | ···················· | ···················· | ···················· | ···················· | ···················· | ···················· |
| U | 3813 hCoV-19/Denmark/DCGC-12501/2020\|EPI_ISL_682682\|202 | ···················· | ···················· | ···················· | ···················· | ···················· | ···················· |
| U | 3814 hCoV-19/Denmark/DCGC-12502/2020\|EPI_ISL_682683\|202 | ···················· | ···················· | ···················· | ···················· | ·····T·············· | ···················· |
| U | 3815 hCoV-19/Denmark/DCGC-12504/2020\|EPI_ISL_682685\|202 | ···················· | ···················· | ···················· | ···················· | ···················· | ···················· |
| U | 3816 hCoV-19/Denmark/DCGC-12506/2020\|EPI_ISL_682687\|202 | ···················· | ·A·················· | ···················· | ···················· | ···················· | ··A················· |
| U | 3817 hCoV-19/Denmark/DCGC-12508/2020\|EPI_ISL_682689\|202 | ···················· | ···················· | ···················· | ···················· | ···················· | ···················· |
| U | 3818 hCoV-19/Denmark/DCGC-12509/2020\|EPI_ISL_682690\|202 | ···················· | ···················· | ···················· | ···················· | ···················· | ···················· |
| U | 3819 hCoV-19/Denmark/DCGC-12510/2020\|EPI_ISL_682691\|202 | ···················· | ·A·················· | ···················· | ···················· | ···················· | ···················· |
| U | 3820 hCoV-19/Denmark/DCGC-12511/2020\|EPI_ISL_682692\|202 | ···················· | ···················· | ···················· | ···················· | ·····T······T······· | ···················· |
| U | 3821 hCoV-19/Denmark/DCGC-12513/2020\|EPI_ISL_682694\|202 | ···················· | ···················· | ···················· | ···················· | ···················· | ···················· |
| U | 3822 hCoV-19/Denmark/DCGC-12515/2020\|EPI_ISL_682696\|202 | ···················· | ···················· | ···················· | ···················· | ···················· | ···················· |
| U | 3823 hCoV-19/Denmark/DCGC-12516/2020\|EPI_ISL_682697\|202 | ···················· | ···················· | ···················· | ···················· | ···················· | ···················· |
| U | 3824 hCoV-19/Denmark/DCGC-12517/2020\|EPI_ISL_682698\|202 | ···················· | ···················· | ···················· | ···················· | ···················· | ···················· |
| U | 3825 hCoV-19/Denmark/DCGC-12518/2020\|EPI_ISL_682699\|202 | ···················· | ···A················ | ···················· | ···················· | ···················· | ···················· |
| U | 3826 hCoV-19/USA/NY-NYCPHL-001361/2020\|EPI_ISL_683982\|2 | ···················· | ···················· | ···················· | ···················· | ···················· | ···················· |
| U | 3827 hCoV-19/USA/NY-NYCPHL-001362/2020\|EPI_ISL_683983\|2 | ···················· | ···················· | ···················· | ···················· | ···················· | ···················· |
| U | 3828 hCoV-19/USA/NY-NYCPHL-001364/2020\|EPI_ISL_683984\|2 | ···················· | ···················· | ···················· | ···················· | ···················· | ···················· |
| U | 3829 hCoV-19/USA/NY-NYCPHL-001368/2020\|EPI_ISL_683985\|2 | ···················· | ···················· | ···················· | ···················· | ···················· | ···················· |
| U | 3830 hCoV-19/USA/NY-NYCPHL-001373/2020\|EPI_ISL_683986\|2 | ···················· | ···················· | ···················· | ···················· | ···················· | ···················· |
| U | 3831 hCoV-19/USA/NY-NYCPHL-001376/2020\|EPI_ISL_683987\|2 | ···················· | ···················· | ···················· | ···················· | ···················· | ···················· |
| U | 3832 hCoV-19/USA/NY-NYCPHL-001378/2020\|EPI_ISL_683988\|2 | ···················· | ·A·················· | ···················· | ···················· | ···················· | ···················· |
| U | 3833 hCoV-19/USA/NY-NYCPHL-001380/2020\|EPI_ISL_683989\|2 | ···················· | ···················· | ···················· | ···················· | ···················· | ···················· |
| U | 3834 hCoV-19/USA/NY-NYCPHL-001384/2020\|EPI_ISL_683990\|2 | ···················· | ···················· | ···················· | ···················· | ···················· | ···················· |
| U | 3835 hCoV-19/USA/NY-NYCPHL-001386/2020\|EPI_ISL_683991\|2 | ···················· | ·A·················· | ···················· | ···················· | ···················· | ···················· |
| U | 3836 hCoV-19/USA/NY-NYCPHL-001387/2020\|EPI_ISL_683992\|2 | ···················· | ···················· | ···················· | ···················· | ···················· | ···················· |
| U | 3837 hCoV-19/USA/NY-NYCPHL-001390/2020\|EPI_ISL_683993\|2 | ···················· | ·A·················· | ···················· | ···················· | ···················· | ···················· |
| U | 3838 hCoV-19/USA/NY-NYCPHL-001392/2020\|EPI_ISL_683994\|2 | ···················· | ···················· | ···················· | ···················· | ···················· | ···················· |
| U | 3839 hCoV-19/USA/NY-NYCPHL-001395/2020\|EPI_ISL_683995\|2 | ···················· | ···················· | ···················· | ···················· | ···················· | ···················· |
| U | 3840 hCoV-19/USA/NY-NYCPHL-001396/2020\|EPI_ISL_683996\|2 | ···················· | ···················· | ···················· | ···················· | ···················· | ···················· |
| U | 3841 hCoV-19/England/MILK-B8D60E/2020\|EPI_ISL_674115\|20 | ···················· | ···················· | ···················· | ···················· | ···················· | ···················· |
| U | 3842 hCoV-19/England/MILK-B8D431/2020\|EPI_ISL_674117\|20 | ···················· | ···················· | ···················· | ···················· | ···················· | ···················· |
| U | 3843 hCoV-19/England/MILK-B86526/2020\|EPI_ISL_674119\|20 | ···················· | ···················· | ···················· | ···················· | ···················· | ···················· |
| U | 3844 hCoV-19/England/MILK-B86632/2020\|EPI_ISL_674124\|20 | ···················· | ···················· | ···················· | ···················· | ···················· | ···················· |
| U | 3845 hCoV-19/England/MILK-B8E4A9/2020\|EPI_ISL_674126\|20 | ···················· | ·A·················· | ···················· | ···················· | ···················· | ··A················· |

| | | | | | | | | |
|---|---|---|---|---|---|---|---|---|
| U | 3846 | hCoV-19/England/MILK-B8684B/2020\|EPI_ISL_674127\|20 | ···················· | ···················· | ···················· | ···················· | ···················· | ···················· |
| U | 3847 | hCoV-19/England/MILK-B86614/2020\|EPI_ISL_674129\|20 | ···················· | ···················· | ···················· | ···················· | ···················· | ···················· |
| U | 3848 | hCoV-19/England/MILK-B8E719/2020\|EPI_ISL_674130\|20 | ···················· | ···················· | ···················· | ···················· | ···················· | ···················· |
| U | 3849 | hCoV-19/England/MILK-B8D54D/2020\|EPI_ISL_674131\|20 | ···················· | ·A·················· | ···················· | ···················· | ···················· | ···A················ |
| U | 3850 | hCoV-19/England/MILK-B8E843/2020\|EPI_ISL_674132\|20 | ···················· | ···················· | ···················· | ···················· | ···················· | ···················· |
| U | 3851 | hCoV-19/England/MILK-B85F74/2020\|EPI_ISL_674134\|20 | ···················· | ···················· | ···················· | ···················· | ······T············· | ···················· |
| U | 3852 | hCoV-19/England/MILK-B8E7CE/2020\|EPI_ISL_674135\|20 | ···················· | ···················· | ···················· | ···················· | ···················· | ···················· |
| U | 3853 | hCoV-19/England/MILK-B8E6EF/2020\|EPI_ISL_674137\|20 | ···················· | ···················· | ···················· | ···················· | ···················· | ···················· |
| U | 3854 | hCoV-19/England/MILK-B8D237/2020\|EPI_ISL_674138\|20 | ···················· | ···················· | ···················· | ···················· | ···················· | ···················· |
| U | 3855 | hCoV-19/England/MILK-B8E5F1/2020\|EPI_ISL_674139\|20 | ···················· | ·A·················· | ···················· | ···················· | ···················· | ···A················ |
| U | 3856 | hCoV-19/England/MILK-B8D3F8/2020\|EPI_ISL_674140\|20 | ···················· | ···················· | ···················· | ···················· | ···················· | ···················· |
| U | 3857 | hCoV-19/England/MILK-B8D2CE/2020\|EPI_ISL_674141\|20 | ···················· | ···················· | ···················· | ···················· | ···················· | ···················· |
| U | 3858 | hCoV-19/England/MILK-B862D4/2020\|EPI_ISL_674142\|20 | ···················· | ···················· | ···················· | ···················· | ···················· | ···················· |
| U | 3859 | hCoV-19/England/MILK-B8E89E/2020\|EPI_ISL_674143\|20 | ···················· | ···················· | ···················· | ···················· | ···················· | ···················· |
| U | 3860 | hCoV-19/England/MILK-B8E746/2020\|EPI_ISL_674145\|20 | ···················· | ···················· | ···················· | ···················· | ···················· | ···················· |
| U | 3861 | hCoV-19/England/MILK-B86702/2020\|EPI_ISL_674147\|20 | ···················· | ···················· | ···················· | ···················· | ···················· | ···················· |
| U | 3862 | hCoV-19/England/MILK-B868B4/2020\|EPI_ISL_674148\|20 | ···················· | ···················· | ···················· | ···················· | ···················· | ···················· |
| U | 3863 | hCoV-19/England/MILK-B85BAD/2020\|EPI_ISL_674149\|20 | ············T······· | ···················· | ···················· | ···················· | ···················· | ···················· |
| U | 3864 | hCoV-19/England/MILK-B863FF/2020\|EPI_ISL_674150\|20 | ···················· | ···················· | ···················· | ···················· | ···················· | ···················· |
| U | 3865 | hCoV-19/England/MILK-B8E728/2020\|EPI_ISL_674151\|20 | ···················· | ···················· | ···················· | ···················· | ···················· | ···················· |
| U | 3866 | hCoV-19/England/MILK-B8E737/2020\|EPI_ISL_674152\|20 | ···················· | ···················· | ···················· | ···················· | ···················· | ···················· |
| U | 3867 | hCoV-19/England/MILK-B8D49B/2020\|EPI_ISL_674153\|20 | ···················· | ···················· | ···················· | ···················· | ···················· | ···················· |
| U | 3868 | hCoV-19/England/MILK-B8681E/2020\|EPI_ISL_674154\|20 | ···················· | ·A·················· | ···················· | ···················· | ···················· | ···A················ |
| U | 3869 | hCoV-19/England/ALDP-B83A0C/2020\|EPI_ISL_674159\|20 | ···················· | ···················· | ···················· | ···················· | ······T············· | ···················· |
| U | 3870 | hCoV-19/England/ALDP-B84C8D/2020\|EPI_ISL_674160\|20 | ···················· | ···················· | ···················· | ···················· | ···················· | ···················· |
| U | 3871 | hCoV-19/England/ALDP-B8440D/2020\|EPI_ISL_674161\|20 | ···················· | ···················· | ···················· | ···················· | ···················· | ···················· |
| U | 3872 | hCoV-19/England/ALDP-B84A74/2020\|EPI_ISL_674162\|20 | ···················· | ···················· | ···················· | ···················· | ···················· | ···················· |
| U | 3873 | hCoV-19/England/ALDP-B84F75/2020\|EPI_ISL_674164\|20 | ···················· | ···················· | ···················· | ···················· | ···················· | ···················· |
| U | 3874 | hCoV-19/England/ALDP-B84F39/2020\|EPI_ISL_674165\|20 | ···················· | ···················· | ···················· | ···················· | ···················· | ···················· |
| U | 3875 | hCoV-19/England/MILK-B8C326/2020\|EPI_ISL_674167\|20 | ···················· | ···················· | ···················· | ···················· | ···················· | ···················· |
| U | 3876 | hCoV-19/England/ALDP-B83DB8/2020\|EPI_ISL_674168\|20 | ···················· | ···················· | ···················· | ···················· | ···················· | ···················· |
| U | 3877 | hCoV-19/England/ALDP-B851F6/2020\|EPI_ISL_674169\|20 | ···················· | ···················· | ···················· | ···················· | ···················· | ···················· |
| U | 3878 | hCoV-19/England/ALDP-B853F0/2020\|EPI_ISL_674170\|20 | ···················· | ···················· | ···················· | ···················· | ···················· | ···················· |
| U | 3879 | hCoV-19/England/ALDP-B85466/2020\|EPI_ISL_674172\|20 | ···················· | ···················· | ···················· | ···················· | ···················· | ···················· |
| U | 3880 | hCoV-19/England/MILK-B8C3CC/2020\|EPI_ISL_674173\|20 | ···················· | ···················· | ···················· | ···················· | ···················· | ···················· |
| U | 3881 | hCoV-19/England/ALDP-B8525D/2020\|EPI_ISL_674174\|20 | ···················· | ···················· | ···················· | ···················· | ···················· | ···················· |
| U | 3882 | hCoV-19/England/ALDP-B84D20/2020\|EPI_ISL_674175\|20 | ···················· | ···················· | ···················· | ···················· | ···················· | ···················· |
| U | 3883 | hCoV-19/England/ALDP-B8527B/2020\|EPI_ISL_674176\|20 | ···················· | ···················· | ···················· | ···················· | ···················· | ···················· |
| U | 3884 | hCoV-19/England/ALDP-B851D8/2020\|EPI_ISL_674177\|20 | ···················· | ···················· | ···················· | ···················· | ···················· | ···················· |
| U | 3885 | hCoV-19/England/MILK-B88524/2020\|EPI_ISL_674178\|20 | ···················· | ···················· | ···················· | ···················· | ···················· | ···················· |
| U | 3886 | hCoV-19/England/ALDP-B85211/2020\|EPI_ISL_674179\|20 | ···················· | ···················· | ···················· | ···················· | ···················· | ···················· |
| U | 3887 | hCoV-19/England/ALDP-B854A2/2020\|EPI_ISL_674181\|20 | ···················· | ···················· | ···················· | ···················· | ···················· | ···A················ |
| U | 3888 | hCoV-19/England/ALDP-B83F94/2020\|EPI_ISL_674182\|20 | ···················· | ·A·················· | ···················· | ···················· | ···················· | ···A················ |
| U | 3889 | hCoV-19/England/MILK-B8C0D4/2020\|EPI_ISL_674183\|20 | ···················· | ···················· | ···················· | ···················· | ···················· | ···A················ |
| U | 3890 | hCoV-19/England/MILK-B8C423/2020\|EPI_ISL_674184\|20 | ···················· | ···················· | ···················· | ···················· | ···················· | ···················· |
| U | 3891 | hCoV-19/England/MILK-B8C4E7/2020\|EPI_ISL_674185\|20 | ···················· | ···················· | ···················· | ···················· | ···················· | ···················· |
| U | 3892 | hCoV-19/England/ALDP-B84CD8/2020\|EPI_ISL_674186\|20 | ···················· | ···················· | ···················· | ···················· | ···················· | ···················· |
| U | 3893 | hCoV-19/England/MILK-B8C308/2020\|EPI_ISL_674187\|20 | ···················· | ···················· | ···················· | ···················· | ···················· | ···················· |
| U | 3894 | hCoV-19/England/ALDP-B84BF9/2020\|EPI_ISL_674188\|20 | ···················· | ···················· | ···················· | ···················· | ···················· | ···················· |
| U | 3895 | hCoV-19/England/MILK-B8C089/2020\|EPI_ISL_674189\|20 | ···················· | ·A·················· | ···················· | ···················· | ···················· | ···A················ |
| U | 3896 | hCoV-19/England/ALDP-B84CC9/2020\|EPI_ISL_674190\|20 | ···················· | ···················· | ···················· | ···················· | ···················· | ···················· |
| U | 3897 | hCoV-19/England/MILK-B8BFC9/2020\|EPI_ISL_674191\|20 | ···················· | ···················· | ···················· | ···················· | ···················· | ···················· |
| U | 3898 | hCoV-19/England/ALDP-B84397/2020\|EPI_ISL_674192\|20 | ···················· | ···················· | ···················· | ···················· | ···················· | ···················· |
| U | 3899 | hCoV-19/England/ALDP-B84A0B/2020\|EPI_ISL_674193\|20 | ···················· | ···················· | ···················· | ···················· | ···················· | ···················· |
| U | 3900 | hCoV-19/England/ALDP-B840BE/2020\|EPI_ISL_674195\|20 | ···················· | ···················· | ···················· | ···················· | ···················· | ···················· |
| U | 3901 | hCoV-19/England/MILK-B8C4D8/2020\|EPI_ISL_674196\|20 | ···················· | ···················· | ···················· | ···················· | ···················· | ···················· |
| U | 3902 | hCoV-19/England/ALDP-B84C23/2020\|EPI_ISL_674197\|20 | ···················· | ···················· | ···················· | ···················· | ···················· | ···················· |
| U | 3903 | hCoV-19/England/MILK-B8C13B/2020\|EPI_ISL_674198\|20 | ···················· | ···················· | ···················· | ···················· | ···················· | ···················· |
| U | 3904 | hCoV-19/England/ALDP-B83F85/2020\|EPI_ISL_674199\|20 | ···················· | ···················· | ···················· | ···················· | ···················· | ···················· |
| U | 3905 | hCoV-19/England/MILK-B88506/2020\|EPI_ISL_674200\|20 | ···················· | ···················· | ···················· | ···················· | ···················· | ···················· |
| U | 3906 | hCoV-19/England/ALDP-B84AFC/2020\|EPI_ISL_674201\|20 | ···················· | ···················· | ···················· | ···················· | ···················· | ···················· |
| U | 3907 | hCoV-19/England/MILK-B881A8/2020\|EPI_ISL_674202\|20 | ···················· | ···················· | ···················· | ···················· | ···················· | ···················· |
| U | 3908 | hCoV-19/England/ALDP-B8434C/2020\|EPI_ISL_674203\|20 | ···················· | ···················· | ···················· | ···················· | ···················· | ···················· |
| U | 3909 | hCoV-19/England/ALDP-B84B44/2020\|EPI_ISL_674204\|20 | ···················· | ···················· | ···················· | ···················· | ···················· | ···················· |
| U | 3910 | hCoV-19/England/MILK-B8C380/2020\|EPI_ISL_674205\|20 | ···················· | ···················· | ···················· | ···················· | ···················· | ···················· |
| U | 3911 | hCoV-19/England/ALDP-B83FA3/2020\|EPI_ISL_674206\|20 | ···················· | ···················· | ···················· | ···················· | ···················· | ···················· |
| U | 3912 | hCoV-19/England/MILK-B88463/2020\|EPI_ISL_674210\|20 | ···················· | ···················· | ···················· | ···················· | ···················· | ···················· |
| U | 3913 | hCoV-19/England/ALDP-B84379/2020\|EPI_ISL_674211\|20 | ···················· | ···················· | ···················· | ···················· | ···················· | ···················· |
| U | 3914 | hCoV-19/England/MILK-B8830C/2020\|EPI_ISL_674212\|20 | ···················· | ···················· | ···················· | ···················· | ···················· | ···················· |
| U | 3915 | hCoV-19/England/MILK-B8C07A/2020\|EPI_ISL_674213\|20 | ···················· | ···················· | ···················· | ···················· | ···················· | ···················· |
| U | 3916 | hCoV-19/England/MILK-B8F271/2020\|EPI_ISL_675446\|20 | ···················· | ···················· | ···················· | ···················· | ···················· | ···················· |
| U | 3917 | hCoV-19/England/ALDP-B8369F/2020\|EPI_ISL_675447\|20 | ···················· | ···················· | ···················· | ···················· | ···················· | ···················· |
| U | 3918 | hCoV-19/England/ALDP-B82F2C/2020\|EPI_ISL_675450\|20 | ···················· | ···················· | ···················· | ···················· | ···················· | ···················· |
| U | 3919 | hCoV-19/England/ALDP-B847C8/2020\|EPI_ISL_675452\|20 | ···················· | ···················· | ···················· | ···················· | ···················· | ···················· |
| U | 3920 | hCoV-19/England/ALDP-B82D31/2020\|EPI_ISL_675454\|20 | ···················· | ···················· | ···················· | ···················· | ·T·················· | ···················· |
| U | 3921 | hCoV-19/England/ALDP-B84537/2020\|EPI_ISL_675455\|20 | ···················· | ···················· | ···················· | ···················· | ···················· | ···················· |
| U | 3922 | hCoV-19/England/ALDP-B84106/2020\|EPI_ISL_675457\|20 | ···················· | ···················· | ···················· | ···················· | ···················· | ···················· |
| U | 3923 | hCoV-19/England/ALDP-B84142/2020\|EPI_ISL_675458\|20 | ···················· | ···················· | ···················· | ···················· | ···················· | ···················· |
| U | 3924 | hCoV-19/England/ALDP-B844D0/2020\|EPI_ISL_675459\|20 | ···················· | ···················· | ···················· | ···················· | ····T·T············· | ···················· |
| U | 3925 | hCoV-19/England/ALDP-B835CF/2020\|EPI_ISL_675460\|20 | ···················· | ···················· | ···················· | ···················· | ···················· | ···················· |

| | | | | | | | | |
|---|---|---|---|---|---|---|---|---|
| U | 3926 | hCoV-19/England/ALDP-B83626/2020\|EPI_ISL_675461\|20 | ···················· | ···················· | ····················· | ···················· | ···················· | ···················· |
| U | 3927 | hCoV-19/England/MILK-B8F165/2020\|EPI_ISL_675462\|20 | ···················· | ···················· | ····················· | ···················· | ···················· | ···················· |
| U | 3928 | hCoV-19/England/ALDP-B82BBF/2020\|EPI_ISL_675464\|20 | ···················· | ···················· | ····················· | ···················· | ···················· | ···················· |
| U | 3929 | hCoV-19/England/ALDP-B8575E/2020\|EPI_ISL_675466\|20 | ···················· | ···················· | ····················· | ···················· | ···················· | ···················· |
| U | 3930 | hCoV-19/England/MILK-B8F38D/2020\|EPI_ISL_675467\|20 | ···················· | ···················· | ····················· | ···················· | ···················· | ···················· |
| U | 3931 | hCoV-19/England/ALDP-B838F3/2020\|EPI_ISL_675468\|20 | ···················· | ···················· | ····················· | ···················· | ·····T·············· | ···················· |
| U | 3932 | hCoV-19/England/ALDP-B842A9/2020\|EPI_ISL_675469\|20 | ···················· | ···················· | ····················· | ···················· | ···················· | ···················· |
| U | 3933 | hCoV-19/England/ALDP-B83AC0/2020\|EPI_ISL_675470\|20 | ···················· | ···················· | ····················· | ···················· | ···················· | ···················· |
| U | 3934 | hCoV-19/England/MILK-B8F0E0/2020\|EPI_ISL_675472\|20 | ···················· | ···················· | ····················· | ···················· | ···················· | ···················· |
| U | 3935 | hCoV-19/England/ALDP-B8377E/2020\|EPI_ISL_675473\|20 | ···················· | ···················· | ····················· | ···················· | ···················· | ···················· |
| U | 3936 | hCoV-19/England/ALDP-B82A2B/2020\|EPI_ISL_675474\|20 | ···················· | ···················· | ····················· | ···················· | ···················· | ···················· |
| U | 3937 | hCoV-19/England/MILK-B8F0B3/2020\|EPI_ISL_675475\|20 | ···················· | ···················· | ····················· | ···················· | ···················· | ···················· |
| U | 3938 | hCoV-19/England/ALDP-B82F59/2020\|EPI_ISL_675476\|20 | ···················· | ···················· | ····················· | ···················· | ···················· | ···················· |
| U | 3939 | hCoV-19/England/ALDP-B83E6A/2020\|EPI_ISL_675477\|20 | ···················· | ···················· | ····················· | ···················· | ···················· | ···················· |
| U | 3940 | hCoV-19/England/ALDP-B834FF/2020\|EPI_ISL_675478\|20 | ···················· | ···················· | ····················· | ···················· | ·····T·············· | ···················· |
| U | 3941 | hCoV-19/England/ALDP-B831CB/2020\|EPI_ISL_675479\|20 | ···················· | ···················· | ····················· | ···················· | ···················· | ···················· |
| U | 3942 | hCoV-19/England/MILK-B89BC7/2020\|EPI_ISL_675480\|20 | ···················· | ···················· | ····················· | ···················· | ···················· | ···················· |
| U | 3943 | hCoV-19/England/ALDP-B82AB2/2020\|EPI_ISL_675481\|20 | ···················· | ···················· | ····················· | ···················· | ···················· | ···················· |
| U | 3944 | hCoV-19/England/ALDP-B831DA/2020\|EPI_ISL_675482\|20 | ···················· | ···················· | ····················· | ···················· | ···················· | ···················· |
| U | 3945 | hCoV-19/England/ALDP-B92703/2020\|EPI_ISL_675483\|20 | ···················· | ···················· | ····················· | ···················· | ···················· | ···················· |
| U | 3946 | hCoV-19/England/ALDP-B83AFD/2020\|EPI_ISL_675484\|20 | ···················· | ···················· | ····················· | ···················· | ···················· | ···················· |
| U | 3947 | hCoV-19/England/MILK-B89EA0/2020\|EPI_ISL_675486\|20 | ···················· | ·A·················· | ····················· | ···················· | ···················· | ··A················· |
| U | 3948 | hCoV-19/England/ALDP-B832E6/2020\|EPI_ISL_675487\|20 | ···················· | ···················· | ····················· | ···················· | ···················· | ···················· |
| U | 3949 | hCoV-19/England/ALDP-B830FB/2020\|EPI_ISL_675488\|20 | ···················· | ···················· | ····················· | ···················· | ···················· | ··A················· |
| U | 3950 | hCoV-19/England/ALDP-B854B1/2020\|EPI_ISL_675489\|20 | ···················· | ···················· | ····················· | ···················· | ···················· | ···················· |
| U | 3951 | hCoV-19/England/ALDP-B9274F/2020\|EPI_ISL_675491\|20 | ···················· | ···················· | ····················· | ···················· | ···················· | ···················· |
| U | 3952 | hCoV-19/England/ALDP-B82FFF/2020\|EPI_ISL_675492\|20 | ···················· | ···················· | ····················· | ···················· | ···················· | ··A················· |
| U | 3953 | hCoV-19/England/ALDP-B82A49/2020\|EPI_ISL_675493\|20 | ···················· | ·A·················· | ····················· | ···················· | ···················· | ··A················· |
| U | 3954 | hCoV-19/England/ALDP-B92466/2020\|EPI_ISL_675494\|20 | ···················· | ···················· | ····················· | ···················· | ···················· | ···················· |
| U | 3955 | hCoV-19/England/MILK-B89E37/2020\|EPI_ISL_675495\|20 | ···················· | ···················· | ····················· | ···················· | ···················· | ···················· |
| U | 3956 | hCoV-19/England/MILK-B89E0A/2020\|EPI_ISL_675496\|20 | ···················· | ···················· | ····················· | ···················· | ···················· | ···················· |
| U | 3957 | hCoV-19/England/ALDP-B83310/2020\|EPI_ISL_675498\|20 | ···················· | ···················· | ····················· | ···················· | ·····T·············· | ···················· |
| U | 3958 | hCoV-19/England/MILK-B89DFE/2020\|EPI_ISL_675499\|20 | ···················· | ···················· | ····················· | ···················· | ···················· | ···················· |
| U | 3959 | hCoV-19/England/MILK-B8BF9C/2020\|EPI_ISL_675500\|20 | ···················· | ···················· | ····················· | ···················· | ···················· | ···················· |
| U | 3960 | hCoV-19/England/ALDP-B8591C/2020\|EPI_ISL_675501\|20 | ···················· | ···················· | ····················· | ···················· | ···················· | ···················· |
| U | 3961 | hCoV-19/England/ALDP-B82AEF/2020\|EPI_ISL_675502\|20 | ···················· | ···················· | ····················· | ···················· | ···················· | ···················· |
| U | 3962 | hCoV-19/England/MILK-B89EDD/2020\|EPI_ISL_675503\|20 | ···················· | ·A·················· | ····················· | ···················· | ···················· | ··A················· |
| U | 3963 | hCoV-19/England/MILK-B89FBC/2020\|EPI_ISL_675504\|20 | ···················· | ···················· | ····················· | ···················· | ···················· | ···················· |
| U | 3964 | hCoV-19/England/MILK-B89E46/2020\|EPI_ISL_675505\|20 | ···················· | ···················· | ····················· | ···················· | ···················· | ···················· |
| U | 3965 | hCoV-19/England/MILK-B89F16/2020\|EPI_ISL_675507\|20 | ···················· | ···················· | ····················· | ···················· | ···················· | ···················· |
| U | 3966 | hCoV-19/England/ALDP-B830EC/2020\|EPI_ISL_675508\|20 | ···················· | ···················· | ····················· | ···················· | ···················· | ···················· |
| U | 3967 | hCoV-19/England/ALDP-B83107/2020\|EPI_ISL_675509\|20 | ···················· | ···················· | ····················· | ···················· | ···················· | ···················· |
| U | 3968 | hCoV-19/England/ALDP-B83222/2020\|EPI_ISL_675510\|20 | ···················· | ···················· | ····················· | ···················· | ···················· | ···················· |
| U | 3969 | hCoV-19/England/ALDP-B8327D/2020\|EPI_ISL_675511\|20 | ···················· | ···················· | ····················· | ···················· | ·····T·············· | ···················· |
| U | 3970 | hCoV-19/England/ALDP-B83028/2020\|EPI_ISL_675512\|20 | ···················· | ···················· | ····················· | ···················· | ···················· | ···················· |
| U | 3971 | hCoV-19/England/MILK-B89E55/2020\|EPI_ISL_675513\|20 | ···················· | ···················· | ····················· | ···················· | ···················· | ···················· |
| U | 3972 | hCoV-19/England/ALDP-B92A73/2020\|EPI_ISL_675514\|20 | ···················· | ···················· | ····················· | ···················· | ···················· | ···················· |
| U | 3973 | hCoV-19/England/ALDP-B8325F/2020\|EPI_ISL_675517\|20 | ···················· | ···················· | ····················· | ···················· | ···················· | ···················· |
| U | 3974 | hCoV-19/England/MILK-B89D85/2020\|EPI_ISL_675518\|20 | ···················· | ···················· | ····················· | ···················· | ···················· | ···················· |
| U | 3975 | hCoV-19/England/ALDP-B831AD/2020\|EPI_ISL_675519\|20 | ···················· | ···················· | ····················· | ···················· | ···················· | ···················· |
| U | 3976 | hCoV-19/England/ALDP-B85B70/2020\|EPI_ISL_675520\|20 | ···················· | ···················· | ····················· | ···················· | ···················· | ···················· |
| U | 3977 | hCoV-19/England/ALDP-B82B28/2020\|EPI_ISL_675521\|20 | ···················· | ·A·················· | ····················· | ···················· | ···················· | ···················· |
| U | 3978 | hCoV-19/England/ALDP-B82B55/2020\|EPI_ISL_675522\|20 | ···················· | ···················· | ····················· | ···················· | ·····T·············· | ···················· |
| U | 3979 | hCoV-19/England/ALDP-B83037/2020\|EPI_ISL_675523\|20 | ···················· | ···················· | ····················· | ···················· | ···················· | ···················· |
| U | 3980 | hCoV-19/England/ALDP-B8592B/2020\|EPI_ISL_675524\|20 | ···················· | ···················· | ····················· | ···················· | ···················· | ···················· |
| U | 3981 | hCoV-19/England/ALDP-B83091/2020\|EPI_ISL_675526\|20 | ···················· | ···················· | ····················· | ···················· | ···················· | ···················· |
| U | 3982 | hCoV-19/England/ALDP-B831BC/2020\|EPI_ISL_675527\|20 | ···················· | ···················· | ····················· | ···················· | ···················· | ···················· |
| U | 3983 | hCoV-19/England/ALDP-B83213/2020\|EPI_ISL_675529\|20 | ···················· | ···················· | ····················· | ···················· | ···················· | ···················· |
| U | 3984 | hCoV-19/England/ALDP-B82EE3/2020\|EPI_ISL_675532\|20 | ···················· | ···················· | ····················· | ···················· | ···················· | ···················· |
| U | 3985 | hCoV-19/England/MILK-B89E19/2020\|EPI_ISL_675533\|20 | ···················· | ···················· | ····················· | ···················· | ···················· | ···················· |
| U | 3986 | hCoV-19/England/ALDP-B830DD/2020\|EPI_ISL_675534\|20 | ···················· | ···················· | ····················· | ···················· | ···················· | ···················· |
| U | 3987 | hCoV-19/England/MILK-B89D67/2020\|EPI_ISL_675535\|20 | ···················· | ···················· | ····················· | ···················· | ···················· | ···················· |
| U | 3988 | hCoV-19/England/ALDP-B82E5C/2020\|EPI_ISL_675536\|20 | ···················· | ·A·················· | ····················· | ···················· | ···················· | ··A················· |
| U | 3989 | hCoV-19/England/ALDP-B925DC/2020\|EPI_ISL_675537\|20 | ···················· | ···················· | ····················· | ···················· | ···················· | ···················· |
| U | 3990 | hCoV-19/England/MILK-B89E91/2020\|EPI_ISL_675538\|20 | ···················· | ···················· | ····················· | ···················· | ···················· | ···················· |
| U | 3991 | hCoV-19/England/ALDP-B85396/2020\|EPI_ISL_675539\|20 | ···················· | ···················· | ····················· | ···················· | ···················· | ···················· |
| U | 3992 | hCoV-19/England/MILK-B89D1C/2020\|EPI_ISL_675542\|20 | ···················· | ···················· | ····················· | ···················· | ···················· | ···················· |
| U | 3993 | hCoV-19/England/ALDP-B852B7/2020\|EPI_ISL_675543\|20 | ···················· | ···················· | ····················· | ···················· | ·····T·············· | ···················· |
| U | 3994 | hCoV-19/England/ALDP-B858C4/2020\|EPI_ISL_675544\|20 | ···················· | ···················· | ····················· | ···················· | ···················· | ···················· |
| U | 3995 | hCoV-19/USA/NY-Wadsworth-31760-01/2020\|EPI_ISL_676 | ···················· | ···················· | ····················· | ···················· | ···················· | ···················· |
| U | 3996 | hCoV-19/USA/NY-Wadsworth-31766-01/2020\|EPI_ISL_676 | ···················· | ···················· | ····················· | ···················· | ···················· | ···················· |
| U | 3997 | hCoV-19/USA/NY-Wadsworth-31786-01/2020\|EPI_ISL_676 | ···················· | ···················· | ····················· | ···················· | ···················· | ···················· |
| U | 3998 | hCoV-19/USA/NY-Wadsworth-32065-01/2020\|EPI_ISL_676 | ···················· | ···················· | ····················· | ···················· | ···················· | ···················· |
| U | 3999 | hCoV-19/USA/NY-Wadsworth-32204-01/2020\|EPI_ISL_676 | ···················· | ···················· | ····················· | ···················· | ···················· | ···················· |
| U | 4000 | hCoV-19/USA/NY-Wadsworth-32317-01/2020\|EPI_ISL_676 | ···················· | ···················· | ····················· | ···················· | ···················· | ···················· |
| U | 4001 | hCoV-19/USA/NY-Wadsworth-32347-01/2020\|EPI_ISL_676 | ···················· | ···················· | ····················· | ···················· | ···················· | ···················· |
| U | 4002 | hCoV-19/USA/NY-Wadsworth-32421-01/2020\|EPI_ISL_676 | ···················· | ···················· | ····················· | ···················· | ···················· | ···················· |
| U | 4003 | hCoV-19/USA/NY-Wadsworth-32670-01/2020\|EPI_ISL_676 | ···················· | ···················· | ····················· | ···················· | ···················· | ···················· |
| U | 4004 | hCoV-19/USA/NY-Wadsworth-32824-01/2020\|EPI_ISL_676 | ···················· | ···················· | ····················· | ···················· | ···················· | ···················· |
| U | 4005 | hCoV-19/USA/NY-Wadsworth-33011-01/2020\|EPI_ISL_676 | ···················· | ···················· | ····················· | ···················· | ···················· | ···················· |

| | | | | | | | |
|---|---|---|---|---|---|---|---|
| U | 4006 hCoV-19/USA/NY-Wadsworth-33026-01/2020\|EPI_ISL_676 | .................... | .................... | ..................... | .................... | .................... | .................... |
| U | 4007 hCoV-19/USA/NY-Wadsworth-33029-01/2020\|EPI_ISL_676 | .................... | .................... | ..................... | .................... | .................... | .................... |
| U | 4008 hCoV-19/USA/NY-Wadsworth-33059-01/2020\|EPI_ISL_676 | .................... | .................... | ..................... | .................... | .................... | .................... |
| U | 4009 hCoV-19/USA/NY-Wadsworth-33126-01/2020\|EPI_ISL_676 | .................... | .................... | ..................... | .................... | .................... | .................... |
| U | 4010 hCoV-19/USA/NY-Wadsworth-33312-01/2020\|EPI_ISL_676 | .................... | .................... | ..................... | .................... | .................... | .................... |
| U | 4011 hCoV-19/USA/NY-Wadsworth-34085-01/2020\|EPI_ISL_676 | .................... | .................... | ..................... | .................... | .................... | .................... |
| U | 4012 hCoV-19/USA/NY-Wadsworth-34564-01/2020\|EPI_ISL_676 | .................... | .................... | ..................... | .................... | .................... | .................... |
| U | 4013 hCoV-19/USA/NY-Wadsworth-34750-01/2020\|EPI_ISL_676 | .................... | .................... | ..................... | .................... | .................... | .................... |
| U | 4014 hCoV-19/USA/NY-Wadsworth-34751-01/2020\|EPI_ISL_676 | .................... | .................... | ..................... | .................... | .................... | .................... |
| U | 4015 hCoV-19/USA/NY-Wadsworth-36550-01/2020\|EPI_ISL_676 | .................... | .................... | ..................... | .................... | .................... | .................... |
| U | 4016 hCoV-19/USA/NY-Wadsworth-37117-01/2020\|EPI_ISL_676 | .................... | .................... | ..................... | .................... | .................... | .................... |
| U | 4017 hCoV-19/USA/NY-Wadsworth-37156-01/2020\|EPI_ISL_676 | .................... | .................... | ..................... | .................... | .................... | .................... |
| U | 4018 hCoV-19/USA/NY-Wadsworth-40148-01/2020\|EPI_ISL_676 | .................... | .................... | ..................... | .................... | .................... | .................... |
| U | 4019 hCoV-19/USA/NY-Wadsworth-40149-01/2020\|EPI_ISL_676 | .................... | .................... | ..................... | .................... | .................... | .................... |
| U | 4020 hCoV-19/USA/NY-Wadsworth-40150-01/2020\|EPI_ISL_676 | .................... | .................... | ..................... | .................... | .................... | .................... |
| U | 4021 hCoV-19/USA/NY-Wadsworth-40156-01/2020\|EPI_ISL_676 | .................... | .................... | ..................... | .................... | .................... | .................... |
| U | 4022 hCoV-19/USA/NY-Wadsworth-40166-01/2020\|EPI_ISL_676 | .................... | .................... | ..................... | .................... | .................... | .................... |
| U | 4023 hCoV-19/USA/NY-Wadsworth-41088-01/2020\|EPI_ISL_676 | .................... | .................... | ..................... | .................... | .................... | .................... |
| U | 4024 hCoV-19/USA/NY-Wadsworth-41091-01/2020\|EPI_ISL_676 | .................... | .................... | ..................... | .................... | .................... | .................... |
| U | 4025 hCoV-19/USA/NY-Wadsworth-41097-01/2020\|EPI_ISL_676 | .................... | .................... | ..................... | .................... | .................... | .................... |
| U | 4026 hCoV-19/USA/NY-Wadsworth-41114-01/2020\|EPI_ISL_676 | .................... | .................... | ..................... | .................... | .................... | .................... |
| U | 4027 hCoV-19/USA/NY-Wadsworth-41115-01/2020\|EPI_ISL_676 | .................... | .................... | ..................... | .................... | .................... | .................... |
| U | 4028 hCoV-19/USA/NY-Wadsworth-41147-01/2020\|EPI_ISL_676 | .................... | .................... | ..................... | .................... | .................... | .................... |
| U | 4029 hCoV-19/USA/NY-Wadsworth-41150-01/2020\|EPI_ISL_676 | .................... | .................... | ..................... | .................... | .................... | .................... |
| U | 4030 hCoV-19/USA/NY-Wadsworth-41193-01/2020\|EPI_ISL_676 | .................... | .................... | ..................... | .................... | .................... | .................... |
| U | 4031 hCoV-19/USA/NY-Wadsworth-41424-01/2020\|EPI_ISL_676 | .................... | .................... | ..................... | .................... | .................... | .................... |
| U | 4032 hCoV-19/USA/NY-Wadsworth-41425-01/2020\|EPI_ISL_676 | .................... | .................... | ..................... | .................... | .................... | .................... |
| U | 4033 hCoV-19/USA/NY-Wadsworth-41428-01/2020\|EPI_ISL_676 | .................... | .................... | ..................... | .................... | .................... | .................... |
| U | 4034 hCoV-19/USA/NY-Wadsworth-41429-01/2020\|EPI_ISL_676 | .................... | .................... | ..................... | .................... | .................... | .................... |
| U | 4035 hCoV-19/USA/NY-Wadsworth-41431-01/2020\|EPI_ISL_676 | .................... | .................... | ..................... | .................... | .................... | .................... |
| U | 4036 hCoV-19/USA/NY-Wadsworth-41432-01/2020\|EPI_ISL_676 | .................... | .................... | ..................... | .................... | .................... | .................... |
| U | 4037 hCoV-19/USA/NY-Wadsworth-41433-01/2020\|EPI_ISL_676 | .................... | .................... | ..................... | .................... | .................... | .................... |
| U | 4038 hCoV-19/USA/NY-Wadsworth-41770-01/2020\|EPI_ISL_676 | .................... | .................... | ..................... | .................... | .................... | .................... |
| U | 4039 hCoV-19/USA/NY-Wadsworth-42494-01/2020\|EPI_ISL_676 | .................... | .................... | ..................... | .................... | .................... | .................... |
| U | 4040 hCoV-19/USA/NY-Wadsworth-43582-01/2020\|EPI_ISL_676 | .................... | .................... | ..................... | .................... | .................... | .................... |
| U | 4041 hCoV-19/USA/NY-Wadsworth-43583-01/2020\|EPI_ISL_676 | .................... | .................... | ..................... | .................... | .................... | .................... |
| U | 4042 hCoV-19/USA/NY-Wadsworth-43613-01/2020\|EPI_ISL_676 | .................... | .................... | ..................... | .................... | .................... | .................... |
| U | 4043 hCoV-19/USA/NY-Wadsworth-43914-01/2020\|EPI_ISL_676 | .................... | .................... | ..................... | .................... | .................... | .................... |
| U | 4044 hCoV-19/USA/NY-Wadsworth-44599-01/2020\|EPI_ISL_676 | .................... | .................... | ..................... | .................... | .................... | .................... |
| U | 4045 hCoV-19/USA/NY-Wadsworth-44602-01/2020\|EPI_ISL_676 | .................... | .................... | ..................... | .................... | .................... | .................... |
| U | 4046 hCoV-19/USA/NY-Wadsworth-44700-01/2020\|EPI_ISL_676 | .................... | .................... | ..................... | .................... | .................... | .................... |
| U | 4047 hCoV-19/USA/NY-Wadsworth-44779-01/2020\|EPI_ISL_676 | .................... | .................... | ..................... | .................... | .................... | .................... |
| U | 4048 hCoV-19/USA/NY-Wadsworth-45314-01/2020\|EPI_ISL_676 | .................... | .................... | ..................... | .................... | .................... | .................... |
| U | 4049 hCoV-19/USA/NY-Wadsworth-46143-01/2020\|EPI_ISL_676 | .................... | .................... | ..................... | .................... | .................... | .................... |
| U | 4050 hCoV-19/USA/NY-Wadsworth-46154-01/2020\|EPI_ISL_676 | .................... | .................... | ..................... | .................... | .................... | .................... |
| U | 4051 hCoV-19/USA/NY-Wadsworth-46181-01/2020\|EPI_ISL_676 | .................... | .................... | ..................... | .................... | .................... | .................... |
| U | 4052 hCoV-19/USA/NY-Wadsworth-46242-01/2020\|EPI_ISL_676 | .................... | .................... | ..................... | .................... | .................... | .................... |
| U | 4053 hCoV-19/USA/NY-Wadsworth-46275-01/2020\|EPI_ISL_676 | .................... | .................... | ..................... | .................... | .................... | .................... |
| U | 4054 hCoV-19/USA/NY-Wadsworth-46358-01/2020\|EPI_ISL_676 | .................... | .................... | ..................... | .................... | .................... | .................... |
| U | 4055 hCoV-19/USA/NY-Wadsworth-46445-01/2020\|EPI_ISL_676 | .................... | .................... | ..................... | .................... | .................... | .................... |
| U | 4056 hCoV-19/USA/NY-Wadsworth-46884-01/2020\|EPI_ISL_676 | .................... | .................... | ..................... | .................... | .................... | .................... |
| U | 4057 hCoV-19/USA/NY-Wadsworth-46906-01/2020\|EPI_ISL_676 | .................... | .................... | ..................... | .................... | .................... | .................... |
| U | 4058 hCoV-19/USA/NY-Wadsworth-46911-01/2020\|EPI_ISL_676 | .................... | .................... | ..................... | .................... | .................... | .................... |
| U | 4059 hCoV-19/USA/NY-Wadsworth-46947-01/2020\|EPI_ISL_676 | .................... | .................... | ..................... | .................... | .................... | .................... |
| U | 4060 hCoV-19/USA/NY-Wadsworth-46957-01/2020\|EPI_ISL_676 | .................... | .................... | ..................... | .................... | .................... | .................... |
| U | 4061 hCoV-19/USA/NY-Wadsworth-46991-01/2020\|EPI_ISL_676 | .................... | .................... | ..................... | .................... | .................... | .................... |
| U | 4062 hCoV-19/USA/NY-Wadsworth-47388-01/2020\|EPI_ISL_676 | .................... | .................... | ..................... | .................... | .................... | .................... |
| U | 4063 hCoV-19/USA/NY-Wadsworth-47399-01/2020\|EPI_ISL_676 | .................... | .................... | ..................... | .................... | .................... | .................... |
| U | 4064 hCoV-19/USA/NY-Wadsworth-47400-01/2020\|EPI_ISL_676 | .................... | .................... | ..................... | .................... | .................... | .................... |
| U | 4065 hCoV-19/USA/NY-Wadsworth-48358-01/2020\|EPI_ISL_676 | .................... | .................... | ..................... | .................... | .................... | .................... |
| U | 4066 hCoV-19/USA/NY-Wadsworth-49019-01/2020\|EPI_ISL_676 | .................... | .................... | ..................... | .................... | .................... | .................... |
| U | 4067 hCoV-19/USA/NY-Wadsworth-50105-01/2020\|EPI_ISL_676 | .................... | .................... | ..................... | .................... | .................... | .................... |
| U | 4068 hCoV-19/USA/NY-Wadsworth-50106-01/2020\|EPI_ISL_676 | .................... | .................... | ..................... | .................... | .................... | .................... |
| U | 4069 hCoV-19/USA/NY-Wadsworth-50110-01/2020\|EPI_ISL_676 | .................... | .................... | ..................... | .................... | .................... | .................... |
| U | 4070 hCoV-19/USA/NY-Wadsworth-50111-01/2020\|EPI_ISL_676 | .................... | .................... | ..................... | .................... | .................... | .................... |
| U | 4071 hCoV-19/USA/NY-Wadsworth-50749-01/2020\|EPI_ISL_676 | .................... | .................... | ..................... | .................... | .................... | .................... |
| U | 4072 hCoV-19/USA/NY-Wadsworth-50892-01/2020\|EPI_ISL_676 | .................... | .................... | ..................... | .................... | .................... | .................... |
| U | 4073 hCoV-19/USA/NY-Wadsworth-51088-01/2020\|EPI_ISL_676 | .................... | .................... | ..................... | .................... | .................... | .................... |
| U | 4074 hCoV-19/USA/NY-Wadsworth-55185-01/2020\|EPI_ISL_676 | .................... | .................... | ..................... | .................... | .................... | .................... |
| U | 4075 hCoV-19/USA/NY-Wadsworth-58806-01/2020\|EPI_ISL_676 | .................... | .................... | ..................... | .................... | .................... | .................... |
| U | 4076 hCoV-19/USA/NY-Wadsworth-58877-01/2020\|EPI_ISL_676 | .................... | .................... | ..................... | .................... | .................... | .................... |
| U | 4077 hCoV-19/USA/NY-Wadsworth-63615-01/2020\|EPI_ISL_676 | .................... | .................... | ..................... | .................... | .................... | .................... |
| U | 4078 hCoV-19/USA/NY-Wadsworth-65664-01/2020\|EPI_ISL_676 | .................... | .................... | ..................... | .................... | .................... | .................... |
| U | 4079 hCoV-19/USA/NY-Wadsworth-65665-01/2020\|EPI_ISL_676 | .................... | .................... | ..................... | .................... | .................... | .................... |
| U | 4080 hCoV-19/USA/NY-Wadsworth-65678-01/2020\|EPI_ISL_676 | .................... | .................... | ..................... | .................... | .................... | .................... |
| U | 4081 hCoV-19/England/BIRM-6A839/2020\|EPI_ISL_679091\|202 | .................... | .................... | ..................... | .................... | .................... | .................... |
| U | 4082 hCoV-19/England/BIRM-6A875/2020\|EPI_ISL_679093\|202 | .................... | .A.................. | ..................... | .................... | .................... | ..A................. |
| U | 4083 hCoV-19/England/BIRM-6A884/2020\|EPI_ISL_679094\|202 | .................... | .................... | ..................... | .................... | .................... | .................... |
| U | 4084 hCoV-19/England/BIRM-6A893/2020\|EPI_ISL_679095\|202 | .................... | .................... | ..................... | .................... | .................... | .................... |
| U | 4085 hCoV-19/England/BIRM-6A8A2/2020\|EPI_ISL_679096\|202 | .................... | .................... | ..................... | .................... | .................... | .................... |

| | | | | | | | | |
|---|---|---|---|---|---|---|---|---|
| U | 4086 hCoV-19/England/BIRM-6A8FD/2020\|EPI_ISL_679098\|202 | ···················· | ···················· | ···················· | ···················· | ···················· | ···················· | |
| U | 4087 hCoV-19/England/BIRM-6A909/2020\|EPI_ISL_679099\|202 | ···················· | ···················· | ···················· | ···················· | ···················· | ···················· | |
| U | 4088 hCoV-19/England/QEUH-B2C759/2020\|EPI_ISL_680078\|20 | ···················· | ···················· | ···················· | ···················· | ···················· | ···················· | |
| U | 4089 hCoV-19/England/QEUH-B2C7A4/2020\|EPI_ISL_680080\|20 | ···················· | ···················· | ···················· | ···················· | ···················· | ···················· | |
| U | 4090 hCoV-19/England/QEUH-B2C7E0/2020\|EPI_ISL_680082\|20 | ···················· | ···················· | ···················· | ···················· | ···················· | ···················· | |
| U | 4091 hCoV-19/England/QEUH-B2C865/2020\|EPI_ISL_680086\|20 | ···················· | ···················· | ···················· | ···················· | ···················· | ···················· | |
| U | 4092 hCoV-19/England/QEUH-B2CFCA/2020\|EPI_ISL_680093\|20 | ···················· | ···················· | ···················· | ···················· | ···················· | ···················· | |
| U | 4093 hCoV-19/England/QEUH-B2CFE8/2020\|EPI_ISL_680094\|20 | ···················· | ···················· | ···················· | ···················· | ···················· | ···················· | |
| U | 4094 hCoV-19/England/QEUH-B2CFF7/2020\|EPI_ISL_680095\|20 | ···················· | ···················· | ···················· | ···················· | ···················· | ···················· | |
| U | 4095 hCoV-19/England/QEUH-B2D002/2020\|EPI_ISL_680096\|20 | ···················· | ···················· | ···················· | ···················· | ···················· | ···················· | |
| U | 4096 hCoV-19/England/QEUH-B2D06C/2020\|EPI_ISL_680098\|20 | ···················· | ···················· | ···················· | ···················· | ···················· | ···················· | |
| U | 4097 hCoV-19/England/MILK-B8E755/2020\|EPI_ISL_674120\|20 | ···················· | ···················· | ···················· | ···················· | ···················· | ···················· | |
| U | 4098 hCoV-19/England/MILK-B86474/2020\|EPI_ISL_674121\|20 | ···················· | ···················· | ···················· | ···················· | ······T············· | ···················· | |
| U | 4099 hCoV-19/England/MILK-B8D3DA/2020\|EPI_ISL_674122\|20 | ···················· | ···················· | ···················· | ···················· | ···················· | ···················· | |
| U | 4100 hCoV-19/England/MILK-B86104/2020\|EPI_ISL_674123\|20 | ···················· | ···················· | ···················· | ···················· | ···················· | ···················· | |
| U | 4101 hCoV-19/England/ALDP-B8395A/2020\|EPI_ISL_675448\|20 | ···················· | ···················· | ···················· | ···················· | ···················· | ···················· | |
| U | 4102 hCoV-19/England/ALDP-B82EF2/2020\|EPI_ISL_675449\|20 | ···················· | ···················· | ···················· | ···················· | ···················· | ···················· | |
| U | 4103 hCoV-19/England/ALDP-B74EA8/2020\|EPI_ISL_679001\|20 | ···················· | ···················· | ···················· | ···················· | ···················· | ···················· | |
| U | 4104 hCoV-19/England/ALDP-B74EB7/2020\|EPI_ISL_679002\|20 | ···················· | ···················· | ···················· | ···················· | ···················· | ···················· | |
| U | 4105 hCoV-19/England/ALDP-B74ED5/2020\|EPI_ISL_679004\|20 | ···················· | ···················· | ···················· | ···················· | ···················· | ···················· | |
| U | 4106 hCoV-19/England/ALDP-B74EE4/2020\|EPI_ISL_679005\|20 | ···················· | ···················· | ···················· | ···················· | ···················· | ···················· | |
| U | 4107 hCoV-19/England/ALDP-B74F0F/2020\|EPI_ISL_679007\|20 | ···················· | ···················· | ···················· | ···················· | ···················· | ···················· | |
| U | 4108 hCoV-19/England/BIRM-69F9F/2020\|EPI_ISL_679009\|202 | ···················· | ···················· | ···················· | ···················· | ···················· | ···················· | |
| U | 4109 hCoV-19/Wales/PHWC-48FC98/2020\|EPI_ISL_681211\|2020 | ···················· | ···················· | ···················· | ···················· | ···················· | ···················· | |
| U | 4110 hCoV-19/Wales/PHWC-48FCA7/2020\|EPI_ISL_681212\|2020 | ···················· | ···················· | ···················· | ···················· | ···················· | ···················· | |
| U | 4114 hCoV-19/Wales/PHWC-48FCF2/2020\|EPI_ISL_681217\|2020 | ···················· | ···················· | ···················· | ···················· | NNN················· | ···················· | 1 |
| U | 4115 hCoV-19/Wales/PHWC-48FD1D/2020\|EPI_ISL_681219\|2020 | ···················· | ···················· | ···················· | ···················· | ···················· | ···················· | |
| U | 4116 hCoV-19/Wales/PHWC-48FD3B/2020\|EPI_ISL_681221\|2020 | ···················· | ···················· | ···················· | ···················· | ···················· | ···················· | |
| U | 4117 hCoV-19/Wales/PHWC-48FD4A/2020\|EPI_ISL_681222\|2020 | ···················· | ···················· | ···················· | ···················· | ···················· | ···················· | |
| U | 4118 hCoV-19/Wales/PHWC-48FD59/2020\|EPI_ISL_681223\|2020 | ···················· | ···················· | ···················· | ···················· | ···················· | ···················· | |
| U | 4119 hCoV-19/Wales/PHWC-48FD77/2020\|EPI_ISL_681225\|2020 | ···················· | ···················· | ···················· | ···················· | ···················· | ···················· | |
| U | 4120 hCoV-19/Wales/PHWC-48FD86/2020\|EPI_ISL_681226\|2020 | ···················· | ···················· | ···················· | ···················· | ···················· | ···················· | |
| U | 4121 hCoV-19/Wales/PHWC-48FD95/2020\|EPI_ISL_681227\|2020 | ···················· | ···················· | ···················· | ···················· | ···················· | ···················· | |
| U | 4122 hCoV-19/Wales/PHWC-48FDB3/2020\|EPI_ISL_681228\|2020 | ···················· | ···················· | ···················· | ···················· | ···················· | ···················· | |
| U | 4124 hCoV-19/Wales/PHWC-48FDD1/2020\|EPI_ISL_681230\|2020 | ···················· | ···················· | ···················· | ···················· | ···················· | ···················· | |
| U | 4126 hCoV-19/Wales/PHWC-48FDFF/2020\|EPI_ISL_681232\|2020 | ···················· | ···················· | ···················· | ···················· | ···················· | ···················· | |
| U | 4127 hCoV-19/Wales/PHWC-48FE0B/2020\|EPI_ISL_681233\|2020 | ···················· | ···················· | ···················· | ···················· | ···················· | ···················· | |
| U | 4129 hCoV-19/Wales/PHWC-48FE38/2020\|EPI_ISL_681236\|2020 | ···················· | ···················· | ···················· | ···················· | ···················· | ···················· | |
| U | 4130 hCoV-19/Wales/PHWC-48FE47/2020\|EPI_ISL_681237\|2020 | ···················· | ···················· | ···················· | ···················· | ···················· | ···················· | |
| U | 4134 hCoV-19/Wales/PHWC-48FEA1/2020\|EPI_ISL_681243\|2020 | ···················· | ···················· | ···················· | ···················· | ···················· | ···················· | |
| U | 4135 hCoV-19/Wales/PHWC-48FEB0/2020\|EPI_ISL_681244\|2020 | ···················· | ···················· | ···················· | ···················· | ···················· | ···················· | |
| U | 4136 hCoV-19/Wales/PHWC-48FECF/2020\|EPI_ISL_681245\|2020 | ···················· | ···················· | ···················· | ···················· | ···················· | ···················· | |
| U | 4138 hCoV-19/Wales/PHWC-48FEFC/2020\|EPI_ISL_681247\|2020 | ···················· | ···················· | ···················· | ···················· | ···················· | ···················· | |
| U | 4140 hCoV-19/Wales/PHWC-48FF26/2020\|EPI_ISL_681250\|2020 | ···················· | ···················· | ···················· | ···················· | ···················· | ···················· | |
| U | 4141 hCoV-19/Wales/PHWC-48FF44/2020\|EPI_ISL_681252\|2020 | ···················· | ···················· | ···················· | ···················· | ···················· | ···················· | |
| U | 4142 hCoV-19/Wales/PHWC-48FF53/2020\|EPI_ISL_681253\|2020 | ···················· | ···················· | ···················· | ···················· | ···················· | ···················· | |
| U | 4143 hCoV-19/Wales/PHWC-48FF62/2020\|EPI_ISL_681254\|2020 | ···················· | ···················· | ···················· | ···················· | ···················· | ···················· | |
| U | 4144 hCoV-19/Wales/PHWC-48FF71/2020\|EPI_ISL_681255\|2020 | ···················· | ···················· | ···················· | ···················· | ···················· | ···················· | |
| U | 4145 hCoV-19/Wales/PHWC-48FF9F/2020\|EPI_ISL_681257\|2020 | ···················· | ···················· | ···················· | ···················· | ···················· | ···················· | |
| U | 4146 hCoV-19/Wales/PHWC-48FFAE/2020\|EPI_ISL_681258\|2020 | ···················· | ···················· | ···················· | ···················· | ···················· | ···················· | |
| U | 4148 hCoV-19/France/ARA-6041/2020\|EPI_ISL_681261\|2020-0 | ···················· | ···················· | ···················· | ···················· | ···················· | ···················· | |
| U | 4149 hCoV-19/France/ARA-6048/2020\|EPI_ISL_681262\|2020-0 | ···················· | ···················· | ···················· | ···················· | ···················· | ···················· | |
| U | 4150 hCoV-19/France/ARA-6245/2020\|EPI_ISL_681263\|2020-0 | ···················· | ···················· | ···················· | ···················· | ···················· | ···················· | |
| U | 4151 hCoV-19/Belgium/ULG-10634/2020\|EPI_ISL_681264\|2020 | ···················· | ···················· | ···················· | ···················· | ···················· | ···················· | |
| U | 4154 hCoV-19/Bahrain/920455398_S12_L001/2020\|EPI_ISL_68 | ···················· | ···················· | ···················· | ···················· | ···················· | ···················· | |
| U | 4155 hCoV-19/Bahrain/920396988_S5_L001/2020\|EPI_ISL_681 | ···················· | ···················· | ···················· | ···················· | ···················· | ···················· | |
| U | 4156 hCoV-19/Bahrain/341007558_S1_L001/2020\|EPI_ISL_681 | ···················· | ···················· | ···················· | ···················· | ···················· | ···················· | |
| U | 4157 hCoV-19/Bahrain/341007184_S4_L001/2020\|EPI_ISL_681 | ···················· | ···················· | ···················· | ···················· | ···················· | ···················· | |
| U | 4158 hCoV-19/Bahrain/340861174_S24_L001/2020\|EPI_ISL_68 | ···················· | ···················· | ···················· | ···················· | ···················· | ···················· | |
| U | 4159 hCoV-19/Bahrain/340858359_S15_L001/2020\|EPI_ISL_68 | ···················· | ···················· | ···················· | ···················· | ···················· | ···················· | |
| U | 4160 hCoV-19/Bahrain/340846550_S17_L001/2020\|EPI_ISL_68 | ···················· | ···················· | ···················· | ···················· | ···················· | ···················· | |
| U | 4161 hCoV-19/Bahrain/340846298_S19_L001/2020\|EPI_ISL_68 | ···················· | ···················· | ···················· | ···················· | ···················· | ···················· | |
| U | 4162 hCoV-19/Bahrain/340958045/2020\|EPI_ISL_681308\|2020 | ···················· | ···················· | ···················· | ···················· | ···················· | ···················· | |
| U | 4163 hCoV-19/Bahrain/340654690/2020\|EPI_ISL_681309\|2020 | ···················· | ···················· | ···················· | ···················· | ···················· | ···················· | |
| U | 4164 hCoV-19/Bahrain/340844843/2020\|EPI_ISL_681310\|2020 | ···················· | ···················· | ···················· | ···················· | ···················· | ···················· | |
| U | 4165 hCoV-19/Bahrain/920374025/2020\|EPI_ISL_681311\|2020 | ···················· | ···················· | ···················· | ···················· | ···················· | ···················· | |
| U | 4166 hCoV-19/Bahrain/920338796/2020\|EPI_ISL_681312\|2020 | ···················· | ···················· | ···················· | ···················· | ···················· | ···················· | |
| U | 4167 hCoV-19/Bahrain/341036784_S6_L001/2020\|EPI_ISL_681 | ···················· | ···················· | ···················· | ···················· | ···················· | ···················· | |
| U | 4168 hCoV-19/Bahrain/341036734_S5_L001/2020\|EPI_ISL_681 | ········T··········· | ···················· | ···················· | ···················· | ···················· | ···················· | |
| U | 4169 hCoV-19/Bahrain/341016882_S17_L001/2020\|EPI_ISL_68 | ···················· | ···················· | ···················· | ···················· | ···················· | ···················· | |
| U | 4170 hCoV-19/Bahrain/341016831_S19_L001/2020\|EPI_ISL_68 | ···················· | ···················· | ···················· | ···················· | ···················· | ···················· | |
| U | 4171 hCoV-19/Bahrain/341016827_S18_L001/2020\|EPI_ISL_68 | ···················· | ···················· | ···················· | ···················· | ···················· | ···················· | |
| U | 4172 hCoV-19/Bahrain/341016006_S14_L001/2020\|EPI_ISL_68 | ···················· | ···················· | ···················· | ···················· | ···················· | ···················· | |
| U | 4173 hCoV-19/Bahrain/341013933_S13_L001/2020\|EPI_ISL_68 | ···········T········ | ···················· | ···················· | ···················· | ···················· | ···················· | |
| U | 4174 hCoV-19/Denmark/DCGC-12357/2020\|EPI_ISL_682538\|202 | ···················· | ···················· | ···················· | ···················· | ···················· | ···················· | |
| U | 4175 hCoV-19/Denmark/DCGC-12359/2020\|EPI_ISL_682540\|202 | ···················· | ···················· | ···················· | ···················· | ···················· | ···················· | |
| U | 4176 hCoV-19/Denmark/DCGC-12360/2020\|EPI_ISL_682541\|202 | ···················· | ···················· | ···················· | ···················· | ···················· | ···················· | |
| U | 4177 hCoV-19/Denmark/DCGC-12362/2020\|EPI_ISL_682543\|202 | ···················· | ···················· | ···················· | ···················· | ···················· | ···················· | |
| U | 4178 hCoV-19/Denmark/DCGC-12364/2020\|EPI_ISL_682545\|202 | ···················· | ···················· | ···················· | ···················· | ···················· | ···················· | |
| U | 4179 hCoV-19/Denmark/DCGC-12365/2020\|EPI_ISL_682546\|202 | ···················· | ···················· | ···················· | ···················· | ···················· | ···················· | |

| | | | | | | | | |
|---|---|---|---|---|---|---|---|---|
| U | 4180 | hCoV-19/Denmark/DCGC-12366/2020\|EPI_ISL_682547\|202 | ···················· | ···················· | ···················· | ···················· | ···················· | ···················· |
| U | 4181 | hCoV-19/Denmark/DCGC-12367/2020\|EPI_ISL_682548\|202 | ···················· | ·A·················· | ···················· | ···················· | ···················· | ···A················ |
| U | 4182 | hCoV-19/Denmark/DCGC-12371/2020\|EPI_ISL_682552\|202 | ···················· | ···················· | ···················· | ···················· | ···················· | ···················· |
| U | 4183 | hCoV-19/Denmark/DCGC-12372/2020\|EPI_ISL_682553\|202 | ···················· | ···················· | ···················· | ···················· | ···················· | ···················· |
| U | 4184 | hCoV-19/Denmark/DCGC-12373/2020\|EPI_ISL_682554\|202 | ···················· | ···················· | ···················· | ···················· | ···················· | ···················· |
| U | 4185 | hCoV-19/Denmark/DCGC-12374/2020\|EPI_ISL_682555\|202 | ···················· | ···················· | ···················· | ···················· | ···················· | ···················· |
| U | 4186 | hCoV-19/Denmark/DCGC-12375/2020\|EPI_ISL_682556\|202 | ···················· | ···················· | ···················· | ···················· | ···················· | ···················· |
| U | 4187 | hCoV-19/Denmark/DCGC-12376/2020\|EPI_ISL_682557\|202 | ···················· | ···················· | ···················· | ···················· | ···················· | ···················· |
| U | 4188 | hCoV-19/Denmark/DCGC-12381/2020\|EPI_ISL_682562\|202 | ···················· | ···················· | ···················· | ···················· | ···················· | ···················· |
| U | 4189 | hCoV-19/Denmark/DCGC-12383/2020\|EPI_ISL_682564\|202 | ···················· | ···················· | ···················· | ···················· | ···················· | ···················· |
| U | 4190 | hCoV-19/Denmark/DCGC-12385/2020\|EPI_ISL_682566\|202 | ···················· | ·A·················· | ···················· | ···················· | ···················· | ···················· |
| U | 4191 | hCoV-19/Denmark/DCGC-12386/2020\|EPI_ISL_682567\|202 | ···················· | ···················· | ···················· | ···················· | ···················· | ···················· |
| U | 4192 | hCoV-19/Denmark/DCGC-12387/2020\|EPI_ISL_682568\|202 | ···················· | ···················· | ···················· | ···················· | ···················· | ···················· |
| U | 4193 | hCoV-19/Denmark/DCGC-12390/2020\|EPI_ISL_682571\|202 | ···················· | ···················· | ···················· | ···················· | ···················· | ···················· |
| U | 4194 | hCoV-19/Denmark/DCGC-12395/2020\|EPI_ISL_682576\|202 | ···················· | ···················· | ···················· | ···················· | ······T············· | ···················· |
| U | 4195 | hCoV-19/Denmark/DCGC-12396/2020\|EPI_ISL_682577\|202 | ···················· | ···················· | ···················· | ···················· | ···················· | ···················· |
| U | 4196 | hCoV-19/Denmark/DCGC-12397/2020\|EPI_ISL_682578\|202 | ···················· | ···················· | ···················· | ···················· | ···················· | ···················· |
| U | 4197 | hCoV-19/Denmark/DCGC-12399/2020\|EPI_ISL_682580\|202 | ···················· | ·A·················· | ···················· | ···················· | ···················· | ···A················ |
| U | 4198 | hCoV-19/Denmark/DCGC-12400/2020\|EPI_ISL_682581\|202 | ···················· | ···················· | ···················· | ···················· | ···················· | ···················· |
| U | 4199 | hCoV-19/Denmark/DCGC-12402/2020\|EPI_ISL_682583\|202 | ···················· | ···················· | ···················· | ···················· | ···················· | ···················· |
| U | 4200 | hCoV-19/Denmark/DCGC-12403/2020\|EPI_ISL_682584\|202 | ···················· | ···················· | ···················· | ···················· | ···················· | ···················· |
| U | 4201 | hCoV-19/Denmark/DCGC-12404/2020\|EPI_ISL_682585\|202 | ···················· | ···················· | ···················· | ···················· | ···················· | ···················· |
| U | 4202 | hCoV-19/Denmark/DCGC-12405/2020\|EPI_ISL_682586\|202 | ···················· | ···················· | ···················· | ···················· | ···················· | ···················· |
| U | 4203 | hCoV-19/Denmark/DCGC-12406/2020\|EPI_ISL_682587\|202 | ···················· | ···················· | ···················· | ···················· | ···················· | ···················· |
| U | 4204 | hCoV-19/Denmark/DCGC-12408/2020\|EPI_ISL_682589\|202 | ···················· | ···················· | ···················· | ···················· | ···················· | ···················· |
| U | 4205 | hCoV-19/Denmark/DCGC-12410/2020\|EPI_ISL_682591\|202 | ···················· | ·A·················· | ···················· | ···················· | ···················· | ···A················ |
| U | 4206 | hCoV-19/Denmark/DCGC-12411/2020\|EPI_ISL_682592\|202 | ···················· | ···················· | ···················· | ···················· | ···················· | ···················· |
| U | 4207 | hCoV-19/Denmark/DCGC-12412/2020\|EPI_ISL_682593\|202 | ···················· | ···················· | ···················· | ···················· | ···················· | ···················· |
| U | 4208 | hCoV-19/Denmark/DCGC-12413/2020\|EPI_ISL_682594\|202 | ···················· | ···················· | ···················· | ···················· | ···················· | ···················· |
| U | 4209 | hCoV-19/Denmark/DCGC-12414/2020\|EPI_ISL_682595\|202 | ···················· | ·A·················· | ···················· | ···················· | ···················· | ···A················ |
| U | 4210 | hCoV-19/Denmark/DCGC-12415/2020\|EPI_ISL_682596\|202 | ···················· | ···················· | ···················· | ···················· | ···················· | ···················· |
| U | 4211 | hCoV-19/Denmark/DCGC-12416/2020\|EPI_ISL_682597\|202 | ···················· | ···················· | ···················· | ···················· | ···················· | ···················· |
| U | 4212 | hCoV-19/Denmark/DCGC-12417/2020\|EPI_ISL_682598\|202 | ···················· | ···················· | ···················· | ···················· | ···················· | ···················· |
| U | 4213 | hCoV-19/Denmark/DCGC-12418/2020\|EPI_ISL_682599\|202 | ···················· | ···················· | ···················· | ···················· | ······T············· | ···················· |
| U | 4214 | hCoV-19/Denmark/DCGC-12419/2020\|EPI_ISL_682600\|202 | ···················· | ···················· | ···················· | ···················· | ···················· | ···················· |
| U | 4215 | hCoV-19/Denmark/DCGC-12420/2020\|EPI_ISL_682601\|202 | ···················· | ···················· | ···················· | ···················· | ···················· | ···················· |
| U | 4216 | hCoV-19/Denmark/DCGC-12423/2020\|EPI_ISL_682604\|202 | ···················· | ···················· | ···················· | ···················· | ···················· | ···················· |
| U | 4217 | hCoV-19/Denmark/DCGC-12424/2020\|EPI_ISL_682605\|202 | ···················· | ···················· | ···················· | ···················· | ···················· | ···················· |
| U | 4218 | hCoV-19/Denmark/DCGC-12425/2020\|EPI_ISL_682606\|202 | ···················· | ···················· | ···················· | ···················· | ···················· | ···················· |
| U | 4219 | hCoV-19/Denmark/DCGC-12426/2020\|EPI_ISL_682607\|202 | ···················· | ···················· | ···················· | ···················· | ···················· | ···················· |
| U | 4220 | hCoV-19/Denmark/DCGC-12427/2020\|EPI_ISL_682608\|202 | ···················· | ···················· | ···················· | ···················· | ···················· | ···················· |
| U | 4221 | hCoV-19/Denmark/DCGC-12428/2020\|EPI_ISL_682609\|202 | ···················· | ····A··············· | ···················· | ···················· | ···················· | ···················· |
| U | 4222 | hCoV-19/Denmark/DCGC-12429/2020\|EPI_ISL_682610\|202 | ···················· | ···················· | ···················· | ···················· | ···················· | ···················· |
| U | 4223 | hCoV-19/Denmark/DCGC-12432/2020\|EPI_ISL_682613\|202 | ···················· | ···················· | ···················· | ···················· | ···················· | ···················· |
| U | 4224 | hCoV-19/Denmark/DCGC-12434/2020\|EPI_ISL_682615\|202 | ···················· | ···················· | ···················· | ···················· | ···················· | ···················· |
| U | 4225 | hCoV-19/Denmark/DCGC-12435/2020\|EPI_ISL_682616\|202 | ···················· | ···················· | ···················· | ···················· | ···················· | ···················· |
| U | 4226 | hCoV-19/Denmark/DCGC-12436/2020\|EPI_ISL_682617\|202 | ···················· | ···················· | ···················· | ···················· | ···················· | ···················· |
| U | 4227 | hCoV-19/Denmark/DCGC-12437/2020\|EPI_ISL_682618\|202 | ···················· | ···················· | ···················· | ···················· | ···················· | ···················· |
| U | 4228 | hCoV-19/Denmark/DCGC-12439/2020\|EPI_ISL_682620\|202 | ···················· | ···················· | ···················· | ···················· | ···················· | ···················· |
| U | 4229 | hCoV-19/Denmark/DCGC-12440/2020\|EPI_ISL_682621\|202 | ···················· | ···················· | ···················· | ···················· | ···················· | ···················· |
| U | 4230 | hCoV-19/Denmark/DCGC-12441/2020\|EPI_ISL_682622\|202 | ···················· | ···················· | ···················· | ···················· | ···················· | ···················· |
| U | 4231 | hCoV-19/Denmark/DCGC-12442/2020\|EPI_ISL_682623\|202 | ···················· | ···················· | ···················· | ···················· | ···················· | ···················· |
| U | 4232 | hCoV-19/Denmark/DCGC-12443/2020\|EPI_ISL_682624\|202 | ···················· | ···················· | ···················· | ···················· | ···················· | ···················· |
| U | 4233 | hCoV-19/Denmark/DCGC-12444/2020\|EPI_ISL_682625\|202 | ···················· | ···················· | ···················· | ···················· | ···················· | ···················· |
| U | 4234 | hCoV-19/Denmark/DCGC-12446/2020\|EPI_ISL_682627\|202 | ···················· | ·A·················· | ···················· | ···················· | ···················· | ···A················ |
| U | 4235 | hCoV-19/Denmark/DCGC-12447/2020\|EPI_ISL_682628\|202 | ···················· | ···················· | ···················· | ···················· | ···················· | ···················· |
| U | 4236 | hCoV-19/Denmark/DCGC-12448/2020\|EPI_ISL_682629\|202 | ···················· | ···················· | ···················· | ···················· | ···················· | ···················· |
| U | 4237 | hCoV-19/Denmark/DCGC-12451/2020\|EPI_ISL_682632\|202 | ···················· | ···················· | ···················· | ···················· | ···················· | ···················· |
| U | 4238 | hCoV-19/Denmark/DCGC-12453/2020\|EPI_ISL_682634\|202 | ···················· | ···················· | ···················· | ···················· | ···················· | ···················· |
| U | 4239 | hCoV-19/Denmark/DCGC-12454/2020\|EPI_ISL_682635\|202 | ···················· | ···················· | ···················· | ···················· | ···················· | ···················· |
| U | 4240 | hCoV-19/Denmark/DCGC-12456/2020\|EPI_ISL_682637\|202 | ···················· | ···················· | ···················· | ···················· | ···················· | ···················· |
| U | 4241 | hCoV-19/Denmark/DCGC-12457/2020\|EPI_ISL_682638\|202 | ···················· | ···················· | ···················· | ···················· | ···················· | ···················· |
| U | 4242 | hCoV-19/Denmark/DCGC-12460/2020\|EPI_ISL_682641\|202 | ···················· | ···················· | ···················· | ···················· | ···················· | ···················· |
| U | 4243 | hCoV-19/Denmark/DCGC-12461/2020\|EPI_ISL_682642\|202 | ···················· | ·A·················· | ···················· | ···················· | ···················· | ···················· |
| U | 4244 | hCoV-19/Denmark/DCGC-12463/2020\|EPI_ISL_682644\|202 | ···················· | ···················· | ···················· | ···················· | ···················· | ···················· |
| U | 4245 | hCoV-19/Denmark/DCGC-12465/2020\|EPI_ISL_682646\|202 | ···················· | ···················· | ···················· | ···················· | ···················· | ···················· |
| U | 4246 | hCoV-19/Denmark/DCGC-12466/2020\|EPI_ISL_682647\|202 | ···················· | ···················· | ···················· | ···················· | ···················· | ···················· |
| U | 4247 | hCoV-19/Denmark/DCGC-12467/2020\|EPI_ISL_682648\|202 | ···················· | ···················· | ···················· | ···················· | ···················· | ···················· |
| U | 4248 | hCoV-19/Denmark/DCGC-12468/2020\|EPI_ISL_682649\|202 | ···················· | ···················· | ···················· | ···················· | ······T············· | ···················· |
| U | 4249 | hCoV-19/USA/NY-NYCPHL-001372/2020\|EPI_ISL_683875\|2 | ···················· | ···················· | ···················· | ···················· | ···················· | ···················· |
| U | 4250 | hCoV-19/USA/NY-NYCPHL-001303/2020\|EPI_ISL_683876\|2 | ···················· | ···················· | ···················· | ···················· | ···················· | ···················· |
| U | 4251 | hCoV-19/USA/NY-NYCPHL-001287/2020\|EPI_ISL_683878\|2 | ···················· | ···················· | ···················· | ···················· | ···················· | ···················· |
| U | 4252 | hCoV-19/USA/NY-NYCPHL-001293/2020\|EPI_ISL_683879\|2 | ···················· | ···················· | ···················· | ···················· | ···················· | ···················· |
| U | 4253 | hCoV-19/USA/NY-NYCPHL-001318/2020\|EPI_ISL_683880\|2 | ···················· | ···················· | ···················· | ···················· | ···················· | ···················· |
| U | 4254 | hCoV-19/USA/NY-NYCPHL-001354/2020\|EPI_ISL_683881\|2 | ···················· | ···················· | ···················· | ···················· | ···················· | ···················· |
| U | 4255 | hCoV-19/USA/NY-NYCPHL-001196/2020\|EPI_ISL_683884\|2 | ···················· | ···················· | ···················· | ···················· | ···················· | ···················· |
| U | 4256 | hCoV-19/USA/NY-NYCPHL-001188/2020\|EPI_ISL_683885\|2 | ···················· | ·A·················· | ···················· | ···················· | ···················· | ···················· |
| U | 4257 | hCoV-19/USA/NY-NYCPHL-001195/2020\|EPI_ISL_683886\|2 | ···················· | ···················· | ···················· | ···················· | ···················· | ···················· |
| U | 4258 | hCoV-19/USA/NY-NYCPHL-001218/2020\|EPI_ISL_683887\|2 | ···················· | ···················· | ···················· | ···················· | ···················· | ···················· |
| U | 4259 | hCoV-19/USA/NY-NYCPHL-001221/2020\|EPI_ISL_683888\|2 | ···················· | ···················· | ···················· | ···················· | ···················· | ···················· |

| | | | | | | | |
|---|---|---|---|---|---|---|---|
| U | 4260 hCoV-19/USA/NY-NYCPHL-001222/2020\|EPI_ISL_683889\|2 | ···················· | ···················· | ····················· | ···················· | ···················· | ···················· |
| U | 4261 hCoV-19/USA/NY-NYCPHL-001226/2020\|EPI_ISL_683890\|2 | ···················· | ···················· | ····················· | ···················· | ···················· | ···················· |
| U | 4262 hCoV-19/USA/NY-NYCPHL-001228/2020\|EPI_ISL_683891\|2 | ···················· | ·A·················· | ····················· | ···················· | ···················· | ···················· |
| U | 4263 hCoV-19/USA/NY-NYCPHL-001230/2020\|EPI_ISL_683892\|2 | ···················· | ···················· | ····················· | ···················· | ···················· | ···················· |
| U | 4264 hCoV-19/USA/NY-NYCPHL-001240/2020\|EPI_ISL_683893\|2 | ···················· | ···················· | ····················· | ···················· | ···················· | ···················· |
| U | 4265 hCoV-19/USA/NY-NYCPHL-001254/2020\|EPI_ISL_683894\|2 | ···················· | ···················· | ····················· | ···················· | ···················· | ···················· |
| U | 4266 hCoV-19/USA/NY-NYCPHL-001256/2020\|EPI_ISL_683895\|2 | ···················· | ···················· | ····················· | ···················· | ···················· | ···················· |
| U | 4267 hCoV-19/USA/NY-NYCPHL-001265/2020\|EPI_ISL_683896\|2 | ···················· | ···················· | ····················· | ···················· | ···················· | ···················· |
| U | 4268 hCoV-19/USA/NY-NYCPHL-001266/2020\|EPI_ISL_683897\|2 | ···················· | ···················· | ····················· | ···················· | ···················· | ···················· |
| U | 4269 hCoV-19/USA/NY-NYCPHL-001267/2020\|EPI_ISL_683898\|2 | ···················· | ···················· | ····················· | ···················· | ···················· | ···················· |
| U | 4270 hCoV-19/USA/NY-NYCPHL-001268/2020\|EPI_ISL_683899\|2 | ···················· | ···················· | ····················· | ···················· | ···················· | ···················· |
| U | 4271 hCoV-19/USA/NY-NYCPHL-001278/2020\|EPI_ISL_683900\|2 | ···················· | ···················· | ····················· | ···················· | ···················· | ···················· |
| U | 4272 hCoV-19/USA/NY-NYCPHL-001284/2020\|EPI_ISL_683901\|2 | ···················· | ···················· | ····················· | ···················· | ···················· | ···················· |
| U | 4273 hCoV-19/USA/NY-NYCPHL-001285/2020\|EPI_ISL_683902\|2 | ···················· | ···················· | ····················· | ···················· | ···················· | ···················· |
| U | 4274 hCoV-19/USA/NY-NYCPHL-001286/2020\|EPI_ISL_683903\|2 | ···················· | ·A·················· | ····················· | ···················· | ···················· | ···················· |
| U | 4275 hCoV-19/USA/NY-NYCPHL-001290/2020\|EPI_ISL_683904\|2 | ···················· | ·A·················· | ····················· | ···················· | ···················· | ···················· |
| U | 4276 hCoV-19/USA/NY-NYCPHL-001295/2020\|EPI_ISL_683905\|2 | ···················· | ···················· | ····················· | ···················· | ···················· | ···················· |
| U | 4277 hCoV-19/USA/NY-NYCPHL-001299/2020\|EPI_ISL_683906\|2 | ···················· | ·A·················· | ····················· | ···················· | ···················· | ···················· |
| U | 4278 hCoV-19/USA/NY-NYCPHL-001300/2020\|EPI_ISL_683907\|2 | ···················· | ·A·················· | ····················· | ···················· | ···················· | ···················· |
| U | 4279 hCoV-19/USA/NY-NYCPHL-001302/2020\|EPI_ISL_683908\|2 | ···················· | ···················· | ····················· | ···················· | ···················· | ···················· |
| U | 4280 hCoV-19/USA/NY-NYCPHL-001304/2020\|EPI_ISL_683909\|2 | ···················· | ·A·················· | ····················· | ···················· | ···················· | ···················· |
| U | 4281 hCoV-19/USA/NY-NYCPHL-001305/2020\|EPI_ISL_683910\|2 | ···················· | ···················· | ····················· | ···················· | ···················· | ···················· |
| U | 4282 hCoV-19/USA/NY-NYCPHL-001307/2020\|EPI_ISL_683911\|2 | ···················· | ···················· | ····················· | ···················· | ···················· | ···················· |
| U | 4283 hCoV-19/USA/NY-NYCPHL-001310/2020\|EPI_ISL_683912\|2 | ···················· | ···················· | ····················· | ···················· | ···················· | ···················· |
| U | 4284 hCoV-19/USA/NY-NYCPHL-001313/2020\|EPI_ISL_683913\|2 | ···················· | ···················· | ····················· | ···················· | ···················· | ···················· |
| U | 4285 hCoV-19/USA/NY-NYCPHL-001314/2020\|EPI_ISL_683914\|2 | ···················· | ···················· | ····················· | ···················· | ···················· | ···················· |
| U | 4286 hCoV-19/USA/NY-NYCPHL-001315/2020\|EPI_ISL_683915\|2 | ···················· | ···················· | ····················· | ···················· | ···················· | ···················· |
| U | 4287 hCoV-19/USA/NY-NYCPHL-001316/2020\|EPI_ISL_683916\|2 | ···················· | ···················· | ····················· | ···················· | ···················· | ···················· |
| U | 4288 hCoV-19/USA/NY-NYCPHL-001319/2020\|EPI_ISL_683917\|2 | ···················· | ···················· | ····················· | ···················· | ···················· | ···················· |
| U | 4289 hCoV-19/USA/NY-NYCPHL-001322/2020\|EPI_ISL_683918\|2 | ···················· | ···················· | ····················· | ···················· | ···················· | ···················· |
| U | 4290 hCoV-19/USA/NY-NYCPHL-001323/2020\|EPI_ISL_683919\|2 | ···················· | ···················· | ····················· | ···················· | ···················· | ···················· |
| U | 4291 hCoV-19/USA/NY-NYCPHL-001326/2020\|EPI_ISL_683920\|2 | ···················· | ·A·················· | ····················· | ···················· | ···················· | ···················· |
| U | 4292 hCoV-19/USA/NY-NYCPHL-001327/2020\|EPI_ISL_683921\|2 | ···················· | ···················· | ····················· | ···················· | ···················· | ···················· |
| U | 4293 hCoV-19/USA/NY-NYCPHL-001328/2020\|EPI_ISL_683922\|2 | ···················· | ···················· | ····················· | ···················· | ···················· | ···················· |
| U | 4294 hCoV-19/USA/NY-NYCPHL-001329/2020\|EPI_ISL_683923\|2 | ···················· | ···················· | ····················· | ···················· | ···················· | ···················· |
| U | 4295 hCoV-19/USA/NY-NYCPHL-001330/2020\|EPI_ISL_683924\|2 | ···················· | ···················· | ····················· | ···················· | ···················· | ···················· |
| U | 4296 hCoV-19/USA/NY-NYCPHL-001339/2020\|EPI_ISL_683925\|2 | ···················· | ···················· | ····················· | ···················· | ···················· | ···················· |
| U | 4297 hCoV-19/USA/NY-NYCPHL-001344/2020\|EPI_ISL_683926\|2 | ···················· | ···················· | ····················· | ···················· | ···················· | ···················· |
| U | 4298 hCoV-19/USA/NY-NYCPHL-001363/2020\|EPI_ISL_683927\|2 | ···················· | ···················· | ····················· | ···················· | ···················· | ···················· |
| U | 4299 hCoV-19/USA/NY-NYCPHL-001366/2020\|EPI_ISL_683928\|2 | ···················· | ···················· | ····················· | ···················· | ···················· | ···················· |
| U | 4300 hCoV-19/USA/NY-NYCPHL-001367/2020\|EPI_ISL_683929\|2 | ···················· | ···················· | ····················· | ···················· | ···················· | ···················· |
| U | 4301 hCoV-19/USA/NY-NYCPHL-001374/2020\|EPI_ISL_683930\|2 | ···················· | ···················· | ····················· | ···················· | ···················· | ···················· |
| U | 4302 hCoV-19/USA/NY-NYCPHL-001375/2020\|EPI_ISL_683931\|2 | ···················· | ···················· | ····················· | ···················· | ···················· | ···················· |
| U | 4303 hCoV-19/USA/NY-NYCPHL-001377/2020\|EPI_ISL_683932\|2 | ···················· | ···················· | ····················· | ···················· | ···················· | ···················· |
| U | 4304 hCoV-19/USA/NY-NYCPHL-001381/2020\|EPI_ISL_683933\|2 | ···················· | ···················· | ····················· | ···················· | ···················· | ···················· |
| U | 4305 hCoV-19/USA/NY-NYCPHL-001382/2020\|EPI_ISL_683934\|2 | ···················· | ···················· | ····················· | ···················· | ···················· | ···················· |
| U | 4306 hCoV-19/USA/NY-NYCPHL-001383/2020\|EPI_ISL_683935\|2 | ···················· | ···················· | ····················· | ···················· | ···················· | ···················· |
| U | 4307 hCoV-19/USA/NY-NYCPHL-001391/2020\|EPI_ISL_683936\|2 | ···················· | ···················· | ····················· | ···················· | ···················· | ···················· |
| U | 4308 hCoV-19/USA/NY-NYCPHL-001193/2020\|EPI_ISL_683937\|2 | ···················· | ·A·················· | ····················· | ···················· | ···················· | ···················· |
| U | 4309 hCoV-19/USA/NY-NYCPHL-001194/2020\|EPI_ISL_683938\|2 | ···················· | ···················· | ····················· | ···················· | ···················· | ···················· |
| U | 4310 hCoV-19/USA/NY-NYCPHL-001231/2020\|EPI_ISL_683939\|2 | ···················· | ·A·················· | ····················· | ···················· | ···················· | ···················· |
| U | 4311 hCoV-19/USA/NY-NYCPHL-001232/2020\|EPI_ISL_683940\|2 | ···················· | ···················· | ····················· | ···················· | ···················· | ···················· |
| U | 4312 hCoV-19/USA/NY-NYCPHL-001233/2020\|EPI_ISL_683941\|2 | ···················· | ···················· | ····················· | ···················· | ···················· | ···················· |
| U | 4313 hCoV-19/USA/NY-NYCPHL-001238/2020\|EPI_ISL_683942\|2 | ···················· | ···················· | ····················· | ···················· | ···················· | ···················· |
| U | 4314 hCoV-19/USA/NY-NYCPHL-001239/2020\|EPI_ISL_683943\|2 | ···················· | ···················· | ····················· | ···················· | ···················· | ···················· |
| U | 4315 hCoV-19/USA/NY-NYCPHL-001242/2020\|EPI_ISL_683944\|2 | ···················· | ···················· | ····················· | ···················· | ···················· | ···················· |
| U | 4316 hCoV-19/USA/NY-NYCPHL-001243/2020\|EPI_ISL_683945\|2 | ···················· | ···················· | ····················· | ···················· | ···················· | ···················· |
| U | 4317 hCoV-19/USA/NY-NYCPHL-001250/2020\|EPI_ISL_683946\|2 | ···················· | ···················· | ····················· | ···················· | ···················· | ···················· |
| U | 4318 hCoV-19/USA/NY-NYCPHL-001251/2020\|EPI_ISL_683947\|2 | ···················· | ···················· | ····················· | ···················· | ···················· | ···················· |
| U | 4319 hCoV-19/USA/NY-NYCPHL-001252/2020\|EPI_ISL_683948\|2 | ···················· | ···················· | ····················· | ···················· | ···················· | ···················· |
| U | 4320 hCoV-19/USA/NY-NYCPHL-001253/2020\|EPI_ISL_683949\|2 | ···················· | ···················· | ····················· | ···················· | ···················· | ···················· |
| U | 4321 hCoV-19/USA/NY-NYCPHL-001255/2020\|EPI_ISL_683950\|2 | ···················· | ···················· | ····················· | ···················· | ···················· | ···················· |
| U | 4322 hCoV-19/USA/NY-NYCPHL-001257/2020\|EPI_ISL_683951\|2 | ···················· | ···················· | ····················· | ···················· | ···················· | ···················· |
| U | 4323 hCoV-19/USA/NY-NYCPHL-001269/2020\|EPI_ISL_683952\|2 | ···················· | ···················· | ····················· | ···················· | ···················· | ···················· |
| U | 4324 hCoV-19/USA/NY-NYCPHL-001272/2020\|EPI_ISL_683953\|2 | ···················· | ···················· | ····················· | ···················· | ···················· | ···················· |
| U | 4325 hCoV-19/USA/NY-NYCPHL-001274/2020\|EPI_ISL_683954\|2 | ···················· | ···················· | ····················· | ···················· | ···················· | ···················· |
| U | 4326 hCoV-19/USA/NY-NYCPHL-001277/2020\|EPI_ISL_683955\|2 | ···················· | ···················· | ····················· | ···················· | ···················· | ···················· |
| U | 4327 hCoV-19/USA/NY-NYCPHL-001283/2020\|EPI_ISL_683956\|2 | ···················· | ·A·················· | ····················· | ···················· | ···················· | ···················· |
| U | 4328 hCoV-19/USA/NY-NYCPHL-001291/2020\|EPI_ISL_683957\|2 | ···················· | ···················· | ····················· | ···················· | ···················· | ···················· |
| U | 4329 hCoV-19/USA/NY-NYCPHL-001292/2020\|EPI_ISL_683958\|2 | ···················· | ·A·················· | ····················· | ···················· | ···················· | ···················· |
| U | 4330 hCoV-19/USA/NY-NYCPHL-001297/2020\|EPI_ISL_683959\|2 | ···················· | ·····A·············· | ····················· | ···················· | ···················· | ···················· |
| U | 4331 hCoV-19/USA/NY-NYCPHL-001301/2020\|EPI_ISL_683960\|2 | ···················· | ···················· | ····················· | ···················· | ···················· | ···················· |
| U | 4332 hCoV-19/USA/NY-NYCPHL-001309/2020\|EPI_ISL_683961\|2 | ···················· | ···················· | ····················· | ···················· | ···················· | ···················· |
| U | 4333 hCoV-19/USA/NY-NYCPHL-001311/2020\|EPI_ISL_683962\|2 | ···················· | ···················· | ····················· | ···················· | ···················· | ···················· |
| U | 4334 hCoV-19/USA/NY-NYCPHL-001312/2020\|EPI_ISL_683963\|2 | ···················· | ·A·················· | ····················· | ···················· | ···················· | ···················· |
| U | 4335 hCoV-19/USA/NY-NYCPHL-001317/2020\|EPI_ISL_683964\|2 | ···················· | ···················· | ····················· | ···················· | ···················· | ···················· |
| U | 4336 hCoV-19/USA/NY-NYCPHL-001320/2020\|EPI_ISL_683965\|2 | ···················· | ·A·················· | ····················· | ···················· | ···················· | ···················· |
| U | 4337 hCoV-19/USA/NY-NYCPHL-001321/2020\|EPI_ISL_683966\|2 | ···················· | ···················· | ····················· | ···················· | ···················· | ···················· |
| U | 4338 hCoV-19/USA/NY-NYCPHL-001325/2020\|EPI_ISL_683967\|2 | ···················· | ···················· | ····················· | ···················· | ···················· | ···················· |
| U | 4339 hCoV-19/USA/NY-NYCPHL-001332/2020\|EPI_ISL_683968\|2 | ···················· | ·A·················· | ····················· | ···················· | ···················· | ···················· |

| | | | | | | | |
|---|---|---|---|---|---|---|---|
| U | 4340 hCoV-19/USA/NY-NYCPHL-001334/2020\|EPI_ISL_683969\|2 | ···················· | ···················· | ···················· | ···················· | ···················· | ···················· |
| U | 4341 hCoV-19/USA/NY-NYCPHL-001335/2020\|EPI_ISL_683970\|2 | ···················· | ···················· | ···················· | ···················· | ···················· | ···················· |
| U | 4342 hCoV-19/USA/NY-NYCPHL-001336/2020\|EPI_ISL_683971\|2 | ···················· | ···················· | ···················· | ···················· | ···················· | ···················· |
| U | 4343 hCoV-19/USA/NY-NYCPHL-001337/2020\|EPI_ISL_683972\|2 | ···················· | ···················· | ···················· | ···················· | ···················· | ···················· |
| U | 4344 hCoV-19/USA/NY-NYCPHL-001338/2020\|EPI_ISL_683973\|2 | ···················· | ···················· | ···················· | ···················· | ···················· | ···················· |
| U | 4345 hCoV-19/USA/NY-NYCPHL-001342/2020\|EPI_ISL_683974\|2 | ···················· | ·A·················· | ···················· | ···················· | ···················· | ···················· |
| U | 4346 hCoV-19/USA/NY-NYCPHL-001345/2020\|EPI_ISL_683975\|2 | ···················· | ·A·················· | ···················· | ···················· | ···················· | ···················· |
| U | 4347 hCoV-19/USA/NY-NYCPHL-001346/2020\|EPI_ISL_683976\|2 | ···················· | ···················· | ···················· | ···················· | ···················· | ···················· |
| U | 4348 hCoV-19/USA/NY-NYCPHL-001350/2020\|EPI_ISL_683977\|2 | ···················· | ···················· | ···················· | ···················· | ···················· | ···················· |
| U | 4349 hCoV-19/USA/NY-NYCPHL-001355/2020\|EPI_ISL_683978\|2 | ···················· | ···················· | ···················· | ···················· | ···················· | ···················· |
| U | 4350 hCoV-19/USA/NY-NYCPHL-001356/2020\|EPI_ISL_683979\|2 | ···················· | ·A·················· | ···················· | ···················· | ···················· | ···················· |
| U | 4351 hCoV-19/USA/NY-NYCPHL-001358/2020\|EPI_ISL_683980\|2 | ···················· | ···················· | ···················· | ···················· | ···················· | ···················· |
| U | 4352 hCoV-19/USA/NY-NYCPHL-001360/2020\|EPI_ISL_683981\|2 | ···················· | ···················· | ···················· | ···················· | ···················· | ···················· |
| U | 4353 hCoV-19/England/MILK-B8E579/2020\|EPI_ISL_674002\|20 | ···················· | ···················· | ···················· | ···················· | ···················· | ···················· |
| U | 4354 hCoV-19/England/MILK-B8E694/2020\|EPI_ISL_674003\|20 | ···················· | ···················· | ···················· | ···················· | ···················· | ···················· |
| U | 4355 hCoV-19/England/MILK-B8E60D/2020\|EPI_ISL_674004\|20 | ···················· | ···················· | ···················· | ···················· | ···················· | ···················· |
| U | 4356 hCoV-19/England/MILK-B8E88F/2020\|EPI_ISL_674007\|20 | ···················· | ···················· | ···················· | ···················· | ···················· | ···················· |
| U | 4357 hCoV-19/England/MILK-B85DD4/2020\|EPI_ISL_674008\|20 | ···················· | ···················· | ···················· | ···················· | ···················· | ···················· |
| U | 4358 hCoV-19/England/MILK-B86580/2020\|EPI_ISL_674009\|20 | ···················· | ···················· | ···················· | ···················· | ········A··········· | ···················· |
| U | 4359 hCoV-19/England/MILK-B860DA/2020\|EPI_ISL_674010\|20 | ···················· | ···················· | ···················· | ···················· | ···················· | ···················· |
| U | 4360 hCoV-19/England/MILK-B8D307/2020\|EPI_ISL_674011\|20 | ···················· | ···················· | ···················· | ···················· | ···················· | ···················· |
| U | 4361 hCoV-19/England/MILK-B8D1D0/2020\|EPI_ISL_674012\|20 | ···················· | ···················· | ···················· | ···················· | ···················· | ···················· |
| U | 4362 hCoV-19/England/MILK-B865BD/2020\|EPI_ISL_674013\|20 | ···················· | ···················· | ···················· | ···················· | ···················· | ···················· |
| U | 4363 hCoV-19/England/MILK-B8D5B6/2020\|EPI_ISL_674014\|20 | ···················· | ···················· | ···················· | ···················· | ···················· | ···················· |
| U | 4364 hCoV-19/England/MILK-B86438/2020\|EPI_ISL_674016\|20 | ···················· | ···················· | ···················· | ···················· | ···················· | ···················· |
| U | 4365 hCoV-19/England/MILK-B8669C/2020\|EPI_ISL_674020\|20 | ···················· | ···················· | ···················· | ···················· | ···················· | ···················· |
| U | 4366 hCoV-19/England/MILK-B8D149/2020\|EPI_ISL_674021\|20 | ···················· | ···················· | ···················· | ···················· | ···················· | ···················· |
| U | 4367 hCoV-19/England/MILK-B8E3AC/2020\|EPI_ISL_674022\|20 | ···················· | ·A·················· | ···················· | ···················· | ···················· | ···A················ |
| U | 4368 hCoV-19/England/MILK-B85DB6/2020\|EPI_ISL_674023\|20 | ···················· | ···················· | ···················· | ···················· | ···················· | ···················· |
| U | 4369 hCoV-19/England/MILK-B8D343/2020\|EPI_ISL_674024\|20 | ···················· | ···················· | ···················· | ···················· | ···················· | ···················· |
| U | 4370 hCoV-19/England/MILK-B86298/2020\|EPI_ISL_674025\|20 | ···················· | ···················· | ···················· | ···················· | ···················· | ···················· |
| U | 4371 hCoV-19/England/MILK-B8E53D/2020\|EPI_ISL_674026\|20 | ···················· | ···················· | ···················· | ···················· | ···················· | ···················· |
| U | 4372 hCoV-19/England/MILK-B8D686/2020\|EPI_ISL_674027\|20 | ···················· | ···················· | ···················· | ···················· | ······T············· | ···················· |
| U | 4373 hCoV-19/England/MILK-B86544/2020\|EPI_ISL_674028\|20 | ···················· | ···················· | ···················· | ···················· | ···················· | ···················· |
| U | 4374 hCoV-19/England/MILK-B86605/2020\|EPI_ISL_674030\|20 | ···················· | ···················· | ···················· | ···················· | ······T············· | ···················· |
| U | 4375 hCoV-19/England/MILK-B8633B/2020\|EPI_ISL_674031\|20 | ···················· | ···················· | ···················· | ···················· | ···················· | ···················· |
| U | 4376 hCoV-19/England/MILK-B85FB0/2020\|EPI_ISL_674032\|20 | ···················· | ···················· | ···················· | ···················· | ···················· | ···················· |
| U | 4377 hCoV-19/England/MILK-B868D2/2020\|EPI_ISL_674033\|20 | ···················· | ···················· | ···················· | ···················· | ······T············· | ···················· |
| U | 4378 hCoV-19/England/MILK-B85ED1/2020\|EPI_ISL_674034\|20 | ···················· | ···················· | ···················· | ···················· | ···················· | ···················· |
| U | 4379 hCoV-19/England/MILK-B861AA/2020\|EPI_ISL_674035\|20 | ···················· | ···················· | ···················· | ···················· | ···················· | ···················· |
| U | 4380 hCoV-19/England/MILK-B86650/2020\|EPI_ISL_674036\|20 | ···················· | ···················· | ···················· | ···················· | ···················· | ···················· |
| U | 4381 hCoV-19/England/MILK-B8E861/2020\|EPI_ISL_674039\|20 | ···················· | ···················· | ···················· | ···················· | ···················· | ···················· |
| U | 4382 hCoV-19/England/MILK-B8D413/2020\|EPI_ISL_674040\|20 | ···················· | ···················· | ···················· | ···················· | ······T············· | ···················· |
| U | 4383 hCoV-19/England/MILK-B8D12B/2020\|EPI_ISL_674041\|20 | ···················· | ···················· | ···················· | ···················· | ···················· | ···················· |
| U | 4384 hCoV-19/England/MILK-B85BF8/2020\|EPI_ISL_674042\|20 | ···················· | ···················· | ···················· | ···················· | ···················· | ···················· |
| U | 4385 hCoV-19/England/MILK-B85E77/2020\|EPI_ISL_674043\|20 | ···················· | ·A·················· | ···················· | ···················· | ···················· | ···A················ |
| U | 4386 hCoV-19/England/MILK-B8D2BF/2020\|EPI_ISL_674044\|20 | ···················· | ···················· | ···················· | ···················· | ···················· | ···················· |
| U | 4387 hCoV-19/England/MILK-B8E412/2020\|EPI_ISL_674045\|20 | ···················· | ···················· | ···················· | ···················· | ···················· | ···················· |
| U | 4388 hCoV-19/England/MILK-B8D246/2020\|EPI_ISL_674046\|20 | ···················· | ···················· | ···················· | ···················· | ···················· | ···················· |
| U | 4389 hCoV-19/England/MILK-B8D158/2020\|EPI_ISL_674047\|20 | ···················· | ·A·················· | ···················· | ···················· | ···················· | ···A················ |
| U | 4390 hCoV-19/England/MILK-B85F29/2020\|EPI_ISL_674048\|20 | ···················· | ···················· | ···················· | ···················· | ···················· | ···················· |
| U | 4391 hCoV-19/England/MILK-B8E6A3/2020\|EPI_ISL_674049\|20 | ···················· | ···················· | ···················· | ···················· | ···················· | ···················· |
| U | 4392 hCoV-19/England/MILK-B85C04/2020\|EPI_ISL_674051\|20 | ···················· | ···················· | ···················· | ···················· | ···················· | ···················· |
| U | 4393 hCoV-19/England/MILK-B8D4F5/2020\|EPI_ISL_674052\|20 | ···················· | ···················· | ···················· | ···················· | ···················· | ···················· |
| U | 4394 hCoV-19/England/MILK-B867F3/2020\|EPI_ISL_674054\|20 | ···················· | ·A·················· | ···················· | ···················· | ···················· | ···A················ |
| U | 4395 hCoV-19/England/MILK-B8D167/2020\|EPI_ISL_674055\|20 | ···················· | ···················· | ···················· | ···················· | ···················· | ···················· |
| U | 4396 hCoV-19/England/MILK-B861C8/2020\|EPI_ISL_674056\|20 | ···················· | ···················· | ···················· | ···················· | ···················· | ···················· |
| U | 4397 hCoV-19/England/MILK-B86623/2020\|EPI_ISL_674057\|20 | ···················· | ···················· | ···················· | ···················· | ···················· | ···················· |
| U | 4398 hCoV-19/England/MILK-B8D589/2020\|EPI_ISL_674058\|20 | ···················· | ···················· | ···················· | ···················· | ···················· | ···················· |
| U | 4399 hCoV-19/England/MILK-B860BC/2020\|EPI_ISL_674059\|20 | ···················· | ···················· | ···················· | ···················· | ···················· | ···················· |
| U | 4400 hCoV-19/England/MILK-B86887/2020\|EPI_ISL_674060\|20 | ···················· | ···················· | ···················· | ···················· | ···················· | ···················· |
| U | 4401 hCoV-19/England/MILK-B8E816/2020\|EPI_ISL_674061\|20 | ···················· | ···················· | ···················· | ···················· | ···················· | ···················· |
| U | 4402 hCoV-19/England/MILK-B85DC5/2020\|EPI_ISL_674063\|20 | ···················· | ···················· | ···················· | ···················· | ···················· | ···················· |
| U | 4403 hCoV-19/England/MILK-B8E8AD/2020\|EPI_ISL_674064\|20 | ···················· | ···················· | ···················· | ···················· | ···················· | ···················· |
| U | 4404 hCoV-19/England/MILK-B8E54C/2020\|EPI_ISL_674065\|20 | ···················· | ·A·················· | ···················· | ···················· | ···················· | ···················· |
| U | 4405 hCoV-19/England/MILK-B8D273/2020\|EPI_ISL_674066\|20 | ···················· | ···················· | ···················· | ···················· | ···················· | ···················· |
| U | 4406 hCoV-19/England/MILK-B868C3/2020\|EPI_ISL_674067\|20 | ···················· | ···················· | ···················· | ···················· | ···················· | ···················· |
| U | 4407 hCoV-19/England/MILK-B8D176/2020\|EPI_ISL_674068\|20 | ···················· | ···················· | ···················· | ···················· | ···················· | ···················· |
| U | 4408 hCoV-19/England/MILK-B8D282/2020\|EPI_ISL_674069\|20 | ···················· | ·A·················· | ···················· | ···················· | ···················· | ···A················ |
| U | 4409 hCoV-19/England/MILK-B85E59/2020\|EPI_ISL_674070\|20 | ···················· | ···················· | ···················· | ···················· | ···················· | ···················· |
| U | 4410 hCoV-19/England/MILK-B868A5/2020\|EPI_ISL_674071\|20 | ···················· | ···················· | ···················· | ···················· | ···················· | ···················· |
| U | 4411 hCoV-19/England/MILK-B8E852/2020\|EPI_ISL_674073\|20 | ···················· | ···················· | ···················· | ···················· | ···················· | ···················· |
| U | 4412 hCoV-19/England/MILK-B85FA1/2020\|EPI_ISL_674074\|20 | ···················· | ···················· | ···················· | ···················· | ···················· | ···················· |
| U | 4413 hCoV-19/England/MILK-B8E667/2020\|EPI_ISL_674075\|20 | ···················· | ···················· | ···················· | ···················· | ···················· | ···················· |
| U | 4414 hCoV-19/England/MILK-B867B7/2020\|EPI_ISL_674076\|20 | ···················· | ···················· | ···················· | ···················· | ···················· | ···················· |
| U | 4415 hCoV-19/England/MILK-B868E1/2020\|EPI_ISL_674077\|20 | ···················· | ·A·················· | ···················· | ···················· | ···················· | ···A················ |
| U | 4416 hCoV-19/England/MILK-B8D668/2020\|EPI_ISL_674079\|20 | ···················· | ···················· | ···················· | ···················· | ···················· | ···················· |
| U | 4417 hCoV-19/England/MILK-B8D71A/2020\|EPI_ISL_674100\|20 | ···················· | ···················· | ···················· | ···················· | ···················· | ···················· |
| U | 4418 hCoV-19/England/MILK-B85EE0/2020\|EPI_ISL_674101\|20 | ···················· | ···················· | ···················· | ···················· | ···················· | ···················· |
| U | 4419 hCoV-19/England/MILK-B8E791/2020\|EPI_ISL_674102\|20 | ···················· | ···················· | ···················· | ···················· | ···················· | ···················· |

| | | | | | | | |
|---|---|---|---|---|---|---|---|
| U | 4420 hCoV-19/England/MILK-B8D5C5/2020\|EPI_ISL_674103\|20 | ···················· | ···················· | ····················· | ···················· | ···················· | ···················· |
| U | 4421 hCoV-19/England/MILK-B8667E/2020\|EPI_ISL_674104\|20 | ···················· | ···················· | ····················· | ···················· | ···················· | ···················· |
| U | 4422 hCoV-19/England/MILK-B86720/2020\|EPI_ISL_674105\|20 | ···················· | ···················· | ····················· | ···················· | ···················· | ···················· |
| U | 4423 hCoV-19/England/MILK-B8D334/2020\|EPI_ISL_674106\|20 | ···················· | ···················· | ····················· | ···················· | ···················· | ···················· |
| U | 4424 hCoV-19/England/MILK-B864FC/2020\|EPI_ISL_674107\|20 | ···················· | ···················· | ····················· | ···················· | ···················· | ···················· |
| U | 4425 hCoV-19/England/MILK-B8D38F/2020\|EPI_ISL_674109\|20 | ···················· | ···················· | ····················· | ···················· | ···················· | ···················· |
| U | 4426 hCoV-19/England/MILK-B8677B/2020\|EPI_ISL_674110\|20 | ···················· | ···················· | ····················· | ···················· | ······T············· | ···················· |
| U | 4427 hCoV-19/England/MILK-B85DA7/2020\|EPI_ISL_674111\|20 | ···················· | ···················· | ····················· | ···················· | ···················· | ···················· |
| U | 4428 hCoV-19/England/MILK-B8D316/2020\|EPI_ISL_674113\|20 | ···················· | ···················· | ····················· | ···················· | ···················· | ···················· |
| U | 4429 hCoV-19/England/MILK-B8D352/2020\|EPI_ISL_674114\|20 | ···················· | ···················· | ····················· | ···················· | ···················· | ···················· |
| U | 4430 hCoV-19/England/ALDP-B83A66/2020\|EPI_ISL_675325\|20 | ···················· | ···················· | ····················· | ···················· | ···················· | ···················· |
| U | 4431 hCoV-19/England/MILK-B8F253/2020\|EPI_ISL_675326\|20 | ···················· | ···················· | ····················· | ···················· | ···················· | ···················· |
| U | 4432 hCoV-19/England/ALDP-B84731/2020\|EPI_ISL_675327\|20 | ···················· | ···················· | ····················· | ···················· | ···················· | ···················· |
| U | 4433 hCoV-19/England/ALDP-B846AD/2020\|EPI_ISL_675330\|20 | ···················· | ···················· | ····················· | ···················· | ···················· | ···················· |
| U | 4434 hCoV-19/England/MILK-B8F262/2020\|EPI_ISL_675331\|20 | ···················· | ···················· | ····················· | ···················· | ···················· | ···················· |
| U | 4435 hCoV-19/England/MILK-B8F235/2020\|EPI_ISL_675332\|20 | ···················· | ···················· | ····················· | ···················· | ···················· | ···················· |
| U | 4436 hCoV-19/England/ALDP-B83AA2/2020\|EPI_ISL_675334\|20 | ···················· | ···················· | ····················· | ···················· | ······T············· | ···················· |
| U | 4437 hCoV-19/England/ALDP-B8428B/2020\|EPI_ISL_675335\|20 | ···················· | ···················· | ····················· | ···················· | ···················· | ···················· |
| U | 4438 hCoV-19/England/ALDP-B85AA0/2020\|EPI_ISL_675336\|20 | ···················· | ···················· | ····················· | ···················· | ···················· | ···················· |
| U | 4439 hCoV-19/England/ALDP-B82BFB/2020\|EPI_ISL_675337\|20 | ···················· | ···················· | ····················· | ···················· | ···················· | ···················· |
| U | 4440 hCoV-19/England/ALDP-B8469E/2020\|EPI_ISL_675339\|20 | ···················· | ···················· | ····················· | ···················· | ···················· | ···················· |
| U | 4441 hCoV-19/England/MILK-B8F350/2020\|EPI_ISL_675340\|20 | ···················· | ···················· | ····················· | ···················· | ···················· | ···················· |
| U | 4442 hCoV-19/England/ALDP-B82F4A/2020\|EPI_ISL_675341\|20 | ···················· | ···················· | ····················· | ···················· | ···················· | ···················· |
| U | 4443 hCoV-19/England/ALDP-B8427C/2020\|EPI_ISL_675343\|20 | ···················· | ···················· | ····················· | ···················· | ···················· | ···················· |
| U | 4444 hCoV-19/England/ALDP-B83723/2020\|EPI_ISL_675344\|20 | ···················· | ···················· | ····················· | ···················· | ···················· | ···················· |
| U | 4445 hCoV-19/England/ALDP-B8492C/2020\|EPI_ISL_675345\|20 | ···················· | ···················· | ····················· | ···················· | ···················· | ···················· |
| U | 4446 hCoV-19/England/ALDP-B84300/2020\|EPI_ISL_675346\|20 | ···················· | ···················· | ····················· | ···················· | ···················· | ···················· |
| U | 4447 hCoV-19/England/ALDP-B85703/2020\|EPI_ISL_675347\|20 | ···················· | ···················· | ····················· | ···················· | ···················· | ···················· |
| U | 4448 hCoV-19/England/ALDP-B845A0/2020\|EPI_ISL_675348\|20 | ···················· | ···················· | ····················· | ···················· | ···················· | ···················· |
| U | 4449 hCoV-19/England/ALDP-B8386C/2020\|EPI_ISL_675349\|20 | ···················· | ···················· | ····················· | ···················· | ···················· | ···················· |
| U | 4450 hCoV-19/England/ALDP-B8384E/2020\|EPI_ISL_675350\|20 | ···················· | ···················· | ····················· | ···················· | ···················· | ···················· |
| U | 4451 hCoV-19/England/ALDP-B83C60/2020\|EPI_ISL_675351\|20 | ···················· | ···················· | ····················· | ···················· | ···················· | ···················· |
| U | 4452 hCoV-19/England/ALDP-B836EA/2020\|EPI_ISL_675352\|20 | ···················· | ···················· | ····················· | ···················· | ···················· | ···················· |
| U | 4453 hCoV-19/England/ALDP-B82F77/2020\|EPI_ISL_675354\|20 | ···················· | ···················· | ····················· | ···················· | ···················· | ···················· |
| U | 4454 hCoV-19/England/ALDP-B83608/2020\|EPI_ISL_675355\|20 | ···················· | ···················· | ····················· | ···················· | ···················· | ···················· |
| U | 4455 hCoV-19/England/ALDP-B83E00/2020\|EPI_ISL_675356\|20 | ···················· | ···················· | ····················· | ···················· | ···················· | ···················· |
| U | 4456 hCoV-19/England/MILK-B8F10B/2020\|EPI_ISL_675357\|20 | ···················· | ···················· | ····················· | ···················· | ···················· | ···················· |
| U | 4457 hCoV-19/England/ALDP-B84670/2020\|EPI_ISL_675358\|20 | ···················· | ···················· | ····················· | ···················· | ···················· | ···················· |
| U | 4458 hCoV-19/England/ALDP-B85ABF/2020\|EPI_ISL_675359\|20 | ···················· | ···················· | ····················· | ···················· | ···················· | ···················· |
| U | 4459 hCoV-19/England/ALDP-B841CA/2020\|EPI_ISL_675360\|20 | ···················· | ···················· | ····················· | ···················· | ···················· | ···················· |
| U | 4460 hCoV-19/England/ALDP-B85721/2020\|EPI_ISL_675361\|20 | ···················· | ···················· | ····················· | ···················· | ···················· | ···················· |
| U | 4461 hCoV-19/England/ALDP-B844FE/2020\|EPI_ISL_675362\|20 | ···················· | ···················· | ····················· | ···················· | ···················· | ···················· |
| U | 4462 hCoV-19/England/ALDP-B85800/2020\|EPI_ISL_675363\|20 | ···················· | ···················· | ····················· | ···················· | ···················· | ···················· |
| U | 4463 hCoV-19/England/MILK-B8EEC9/2020\|EPI_ISL_675364\|20 | ···················· | ···················· | ····················· | ···················· | ···················· | ···················· |
| U | 4464 hCoV-19/England/ALDP-B83E88/2020\|EPI_ISL_675366\|20 | ···················· | ···················· | ····················· | ···················· | ······T············· | ···················· |
| U | 4465 hCoV-19/England/ALDP-B82F1D/2020\|EPI_ISL_675367\|20 | ···················· | ···················· | ····················· | ···················· | ···················· | ···················· |
| U | 4466 hCoV-19/England/MILK-B8F3F6/2020\|EPI_ISL_675368\|20 | ···················· | ···················· | ····················· | ···················· | ······T············· | ···················· |
| U | 4467 hCoV-19/England/MILK-B8EF99/2020\|EPI_ISL_675371\|20 | ···················· | ···················· | ····················· | ···················· | ···················· | ···················· |
| U | 4468 hCoV-19/England/ALDP-B835FC/2020\|EPI_ISL_675373\|20 | ···················· | ···················· | ····················· | ···················· | ···················· | ···················· |
| U | 4469 hCoV-19/England/ALDP-B845FB/2020\|EPI_ISL_675374\|20 | ···················· | ···················· | ····················· | ···················· | ···················· | ···················· |
| U | 4470 hCoV-19/England/ALDP-B82D5F/2020\|EPI_ISL_675375\|20 | ···················· | ···················· | ····················· | ···················· | ···················· | ···················· |
| U | 4471 hCoV-19/England/ALDP-B846DA/2020\|EPI_ISL_675376\|20 | ···················· | ···················· | ····················· | ···················· | ···················· | ···················· |
| U | 4472 hCoV-19/England/ALDP-B8417F/2020\|EPI_ISL_675377\|20 | ···················· | ···················· | ····················· | ···················· | ···················· | ···················· |
| U | 4473 hCoV-19/England/ALDP-B84652/2020\|EPI_ISL_675378\|20 | ···················· | ···················· | ····················· | ···················· | ···················· | ···················· |
| U | 4474 hCoV-19/England/ALDP-B8390F/2020\|EPI_ISL_675380\|20 | ···················· | ···················· | ····················· | ···················· | ···················· | ···················· |
| U | 4475 hCoV-19/England/ALDP-B8383F/2020\|EPI_ISL_675381\|20 | ···················· | ···················· | ····················· | ···················· | ···················· | ···················· |
| U | 4476 hCoV-19/England/ALDP-B847E6/2020\|EPI_ISL_675382\|20 | ···················· | ···················· | ····················· | ···················· | ···················· | ···················· |
| U | 4477 hCoV-19/England/ALDP-B851BA/2020\|EPI_ISL_675383\|20 | ···················· | ···················· | ····················· | ···················· | ···················· | ···················· |
| U | 4478 hCoV-19/England/ALDP-B84124/2020\|EPI_ISL_675385\|20 | ···················· | ···················· | ····················· | ···················· | ···················· | ···················· |
| U | 4479 hCoV-19/England/ALDP-B8429A/2020\|EPI_ISL_675387\|20 | ···················· | ···················· | ····················· | ···················· | ···················· | ···················· |
| U | 4480 hCoV-19/England/ALDP-B84133/2020\|EPI_ISL_675388\|20 | ···················· | ···················· | ····················· | ···················· | ···················· | ···················· |
| U | 4481 hCoV-19/England/MILK-B8F420/2020\|EPI_ISL_675389\|20 | ···················· | ···················· | ····················· | ···················· | ···················· | ···················· |
| U | 4482 hCoV-19/England/ALDP-B856E8/2020\|EPI_ISL_675390\|20 | ···················· | ···················· | ····················· | ···················· | ···················· | ···················· |
| U | 4483 hCoV-19/England/ALDP-B84221/2020\|EPI_ISL_675394\|20 | ···················· | ···················· | ····················· | ···················· | ······T············· | ···················· |
| U | 4484 hCoV-19/England/ALDP-B845EC/2020\|EPI_ISL_675395\|20 | ···················· | ···················· | ····················· | ···················· | ···················· | ···················· |
| U | 4485 hCoV-19/England/MILK-B8F3D8/2020\|EPI_ISL_675397\|20 | ···················· | ···················· | ····················· | ···················· | ···················· | ···················· |
| U | 4486 hCoV-19/England/ALDP-B84977/2020\|EPI_ISL_675398\|20 | ···················· | ···················· | ····················· | ···················· | ···················· | ···················· |
| U | 4487 hCoV-19/England/MILK-B8F03B/2020\|EPI_ISL_675399\|20 | ···················· | ···················· | ····················· | ···················· | ···················· | ···················· |
| U | 4488 hCoV-19/England/ALDP-B8432E/2020\|EPI_ISL_675400\|20 | ···················· | ···················· | ····················· | ···················· | ···················· | ···················· |
| U | 4489 hCoV-19/England/ALDP-B82D40/2020\|EPI_ISL_675401\|20 | ···················· | ···················· | ····················· | ···················· | ···················· | ···················· |
| U | 4490 hCoV-19/England/ALDP-B8376F/2020\|EPI_ISL_675402\|20 | ···················· | ···················· | ····················· | ···················· | ···················· | ···················· |
| U | 4491 hCoV-19/England/ALDP-B857A9/2020\|EPI_ISL_675403\|20 | ···················· | ···················· | ····················· | ···················· | ···················· | ···················· |
| U | 4492 hCoV-19/England/MILK-B8F138/2020\|EPI_ISL_675404\|20 | ···················· | ···················· | ····················· | ···················· | ···················· | ···················· |
| U | 4493 hCoV-19/England/ALDP-B83705/2020\|EPI_ISL_675405\|20 | ···················· | ···················· | ····················· | ···················· | ···················· | ···················· |
| U | 4494 hCoV-19/England/ALDP-B845DD/2020\|EPI_ISL_675407\|20 | ···················· | ···················· | ····················· | ···················· | ···················· | ···················· |
| U | 4495 hCoV-19/England/ALDP-B82D7D/2020\|EPI_ISL_675408\|20 | ···················· | ···················· | ····················· | ···················· | ···················· | ···················· |
| U | 4496 hCoV-19/England/ALDP-B85879/2020\|EPI_ISL_675410\|20 | ···················· | ···················· | ····················· | ···················· | ···················· | ···················· |
| U | 4497 hCoV-19/England/ALDP-B84713/2020\|EPI_ISL_675411\|20 | ···················· | ···················· | ····················· | ···················· | ···················· | ···················· |
| U | 4498 hCoV-19/England/ALDP-B846F8/2020\|EPI_ISL_675412\|20 | ···················· | ···················· | ····················· | ···················· | ···················· | ···················· |
| U | 4499 hCoV-19/England/ALDP-B857E5/2020\|EPI_ISL_675414\|20 | ···················· | ···················· | ····················· | ···················· | ···················· | ···················· |

| | | | | | | | | |
|---|---|---|---|---|---|---|---|---|
| U | 4500 | hCoV-19/England/ALDP-B83AEE/2020\|EPI_ISL_675415\|20 | ···················· | ···················· | ···················· | ···················· | ···················· | ···················· |
| U | 4501 | hCoV-19/England/ALDP-B84968/2020\|EPI_ISL_675416\|20 | ···················· | ···················· | ···················· | ···················· | ···················· | ···················· |
| U | 4502 | hCoV-19/England/MILK-B8F208/2020\|EPI_ISL_675417\|20 | ···················· | ···················· | ···················· | ···················· | ···················· | ···················· |
| U | 4503 | hCoV-19/England/ALDP-B83CD9/2020\|EPI_ISL_675418\|20 | ···················· | ···················· | ···················· | ···················· | ···················· | ···················· |
| U | 4504 | hCoV-19/England/MILK-B8EFE4/2020\|EPI_ISL_675420\|20 | ···················· | ···················· | ···················· | ···················· | ···················· | ···················· |
| U | 4505 | hCoV-19/England/MILK-B8EE7E/2020\|EPI_ISL_675423\|20 | ···················· | ···················· | ···················· | ···················· | ···················· | ···················· |
| U | 4506 | hCoV-19/England/MILK-B8F341/2020\|EPI_ISL_675424\|20 | ···················· | ···················· | ···················· | ···················· | ···················· | ···················· |
| U | 4507 | hCoV-19/England/ALDP-B8494A/2020\|EPI_ISL_675425\|20 | ···················· | ···················· | ···················· | ···················· | ···················· | ···················· |
| U | 4508 | hCoV-19/England/ALDP-B836AE/2020\|EPI_ISL_675426\|20 | ···················· | ···················· | ···················· | ···················· | ···················· | ···················· |
| U | 4509 | hCoV-19/England/ALDP-B84528/2020\|EPI_ISL_675427\|20 | ···················· | ···················· | ···················· | ···················· | ···················· | ···················· |
| U | 4510 | hCoV-19/England/ALDP-B829D3/2020\|EPI_ISL_675428\|20 | ···················· | ···················· | ···················· | ···················· | ···················· | ···················· |
| U | 4511 | hCoV-19/England/ALDP-B84661/2020\|EPI_ISL_675430\|20 | ···················· | ·A·················· | ···················· | ···················· | ···················· | ··A················· |
| U | 4512 | hCoV-19/England/MILK-B8F095/2020\|EPI_ISL_675432\|20 | ···················· | ···················· | ···················· | ···················· | ···················· | ···················· |
| U | 4513 | hCoV-19/England/ALDP-B83CBB/2020\|EPI_ISL_675433\|20 | ···················· | ···················· | ···················· | ···················· | ···················· | ···················· |
| U | 4514 | hCoV-19/England/ALDP-B841AC/2020\|EPI_ISL_675434\|20 | ···················· | ···················· | ···················· | ···················· | ···················· | ···················· |
| U | 4515 | hCoV-19/England/ALDP-B84494/2020\|EPI_ISL_675435\|20 | ···················· | ···················· | ···················· | ···················· | ···················· | ···················· |
| U | 4516 | hCoV-19/England/MILK-B8EFF3/2020\|EPI_ISL_675436\|20 | ···················· | ···················· | ···················· | ···················· | ···················· | ···················· |
| U | 4517 | hCoV-19/England/ALDP-B857B8/2020\|EPI_ISL_675437\|20 | ···················· | ···················· | ···················· | ···················· | ···················· | ···················· |
| U | 4518 | hCoV-19/England/ALDP-B8391E/2020\|EPI_ISL_675438\|20 | ···················· | ···················· | ···················· | ···················· | ···················· | ···················· |
| U | 4519 | hCoV-19/England/MILK-B8EF4E/2020\|EPI_ISL_675439\|20 | ···················· | ···················· | ···················· | ···················· | ···················· | ···················· |
| U | 4520 | hCoV-19/England/ALDP-B83FC1/2020\|EPI_ISL_675440\|20 | ···················· | ·········T·········· | ···················· | ···················· | ···················· | ···················· |
| U | 4521 | hCoV-19/England/ALDP-B83EC4/2020\|EPI_ISL_675441\|20 | ···················· | ···················· | ···················· | ···················· | ···················· | ···················· |
| U | 4522 | hCoV-19/England/ALDP-B8426D/2020\|EPI_ISL_675442\|20 | ···················· | ···················· | ···················· | ···················· | ···················· | ···················· |
| U | 4523 | hCoV-19/England/MILK-B8F0D1/2020\|EPI_ISL_675443\|20 | ···················· | ···················· | ···················· | ···················· | ···················· | ···················· |
| U | 4524 | hCoV-19/England/MILK-B8F1ED/2020\|EPI_ISL_675444\|20 | ···················· | ···················· | ···················· | ···················· | ···················· | ···················· |
| U | 4525 | hCoV-19/England/ALDP-B83662/2020\|EPI_ISL_675445\|20 | ···················· | ···················· | ···················· | ···················· | ···················· | ···················· |
| U | 4526 | hCoV-19/USA/NY-Wadsworth-69881-01/2020\|EPI_ISL_676 | ···················· | ···················· | ···················· | ···················· | ···················· | ···················· |
| U | 4527 | hCoV-19/USA/NY-Wadsworth-96980-01/2020\|EPI_ISL_676 | ···················· | ···················· | ···················· | ···················· | ···················· | ···················· |
| U | 4528 | hCoV-19/USA/NY-Wadsworth-101072-01/2020\|EPI_ISL_67 | ···················· | ···················· | ···················· | ···················· | ···················· | ···················· |
| U | 4529 | hCoV-19/USA/NY-Wadsworth-84414-01/2020\|EPI_ISL_676 | ···················· | ···················· | ···················· | ···················· | ···················· | ···················· |
| U | 4530 | hCoV-19/USA/NY-Wadsworth-10493-02/2020\|EPI_ISL_676 | ···················· | ···················· | ···················· | ···················· | ···················· | ···················· |
| U | 4531 | hCoV-19/USA/NY-Wadsworth-11086-01/2020\|EPI_ISL_676 | ···················· | ···················· | ···················· | ···················· | ···················· | ···················· |
| U | 4532 | hCoV-19/USA/NY-Wadsworth-11205-01/2020\|EPI_ISL_676 | ···················· | ···················· | ···················· | ···················· | ···················· | ···················· |
| U | 4533 | hCoV-19/USA/NY-Wadsworth-12437-01/2020\|EPI_ISL_676 | ···················· | ···················· | ···················· | ···················· | ···················· | ···················· |
| U | 4534 | hCoV-19/USA/NY-Wadsworth-013081-01/2020\|EPI_ISL_67 | ···················· | ···················· | ···················· | ···················· | ···················· | ···················· |
| U | 4535 | hCoV-19/USA/NY-Wadsworth-13288-01/2020\|EPI_ISL_676 | ···················· | ···················· | ···················· | ···················· | ···················· | ···················· |
| U | 4536 | hCoV-19/USA/NY-Wadsworth-14186-01/2020\|EPI_ISL_676 | ···················· | ···················· | ···················· | ···················· | ···················· | ···················· |
| U | 4537 | hCoV-19/USA/NY-Wadsworth-14380-01/2020\|EPI_ISL_676 | ···················· | ···················· | ···················· | ···················· | ···················· | ···················· |
| U | 4538 | hCoV-19/USA/NY-Wadsworth-14427-04/2020\|EPI_ISL_676 | ···················· | ···················· | ···················· | ···················· | ···················· | ···················· |
| U | 4539 | hCoV-19/USA/NY-Wadsworth-14524-01/2020\|EPI_ISL_676 | ···················· | ···················· | ···················· | ···················· | ···················· | ···················· |
| U | 4540 | hCoV-19/USA/NY-Wadsworth-16084-01/2020\|EPI_ISL_676 | ···················· | ···················· | ···················· | ···················· | ···················· | ···················· |
| U | 4541 | hCoV-19/USA/NY-Wadsworth-16115-01/2020\|EPI_ISL_676 | ···················· | ···················· | ···················· | ···················· | ···················· | ···················· |
| U | 4542 | hCoV-19/USA/NY-Wadsworth-16175-01/2020\|EPI_ISL_676 | ···················· | ···················· | ···················· | ···················· | ···················· | ···················· |
| U | 4543 | hCoV-19/USA/NY-Wadsworth-16217-02/2020\|EPI_ISL_676 | ···················· | ···················· | ···················· | ···················· | ···················· | ···················· |
| U | 4544 | hCoV-19/USA/NY-Wadsworth-16555-01/2020\|EPI_ISL_676 | ···················· | ···················· | ···················· | ···················· | ···················· | ···················· |
| U | 4545 | hCoV-19/USA/NY-Wadsworth-16600-01/2020\|EPI_ISL_676 | ···················· | ···················· | ···················· | ···················· | ···················· | ···················· |
| U | 4546 | hCoV-19/USA/NY-Wadsworth-18124-01/2020\|EPI_ISL_676 | ···················· | ···················· | ···················· | ···················· | ···················· | ···················· |
| U | 4547 | hCoV-19/USA/NY-Wadsworth-18534-01/2020\|EPI_ISL_676 | ···················· | ···················· | ···················· | ···················· | ···················· | ···················· |
| U | 4548 | hCoV-19/USA/NY-Wadsworth-18758-01/2020\|EPI_ISL_676 | ···················· | ···················· | ···················· | ···················· | ···················· | ···················· |
| U | 4549 | hCoV-19/USA/NY-Wadsworth-20071-01/2020\|EPI_ISL_676 | ···················· | ···················· | ···················· | ···················· | ···················· | ···················· |
| U | 4550 | hCoV-19/USA/NY-Wadsworth-20843-01/2020\|EPI_ISL_676 | ···················· | ···················· | ···················· | ···················· | ···················· | ···················· |
| U | 4551 | hCoV-19/USA/NY-Wadsworth-22016-01/2020\|EPI_ISL_676 | ···················· | ···················· | ···················· | ···················· | ···················· | ···················· |
| U | 4552 | hCoV-19/USA/NY-Wadsworth-22502-01/2020\|EPI_ISL_676 | ···················· | ···················· | ···················· | ···················· | ···················· | ···················· |
| U | 4553 | hCoV-19/USA/NY-Wadsworth-22514-01/2020\|EPI_ISL_676 | ···················· | ···················· | ···················· | ···················· | ···················· | ···················· |
| U | 4554 | hCoV-19/USA/NY-Wadsworth-23321-01/2020\|EPI_ISL_676 | ···················· | ···················· | ···················· | ···················· | ···················· | ···················· |
| U | 4555 | hCoV-19/USA/NY-Wadsworth-24179-01/2020\|EPI_ISL_676 | ···················· | ···················· | ···················· | ···················· | ···················· | ···················· |
| U | 4556 | hCoV-19/USA/NY-Wadsworth-24215-01/2020\|EPI_ISL_676 | ···················· | ···················· | ···················· | ···················· | ···················· | ···················· |
| U | 4557 | hCoV-19/USA/NY-Wadsworth-24216-01/2020\|EPI_ISL_676 | ···················· | ···················· | ···················· | ···················· | ···················· | ···················· |
| U | 4558 | hCoV-19/USA/NY-Wadsworth-24219-01/2020\|EPI_ISL_676 | ···················· | ···················· | ···················· | ···················· | ···················· | ···················· |
| U | 4559 | hCoV-19/USA/NY-Wadsworth-25299-01/2020\|EPI_ISL_676 | ···················· | ···················· | ···················· | ···················· | ···················· | ···················· |
| U | 4560 | hCoV-19/USA/NY-Wadsworth-25410-01/2020\|EPI_ISL_676 | ···················· | ···················· | ···················· | ···················· | ···················· | ···················· |
| U | 4561 | hCoV-19/USA/NY-Wadsworth-27235-01/2020\|EPI_ISL_676 | ···················· | ···················· | ···················· | ···················· | ···················· | ···················· |
| U | 4562 | hCoV-19/USA/NY-Wadsworth-27271-01/2020\|EPI_ISL_676 | ···················· | ···················· | ···················· | ···················· | ···················· | ···················· |
| U | 4563 | hCoV-19/USA/NY-Wadsworth-27896-01/2020\|EPI_ISL_676 | ···················· | ···················· | ···················· | ···················· | ···················· | ···················· |
| U | 4564 | hCoV-19/USA/NY-Wadsworth-28993-01/2020\|EPI_ISL_676 | ···················· | ···················· | ···················· | ···················· | ···················· | ···················· |
| U | 4565 | hCoV-19/USA/NY-Wadsworth-29118-01/2020\|EPI_ISL_676 | ···················· | ···················· | ···················· | ···················· | ···················· | ···················· |
| U | 4566 | hCoV-19/USA/NY-Wadsworth-29242-01/2020\|EPI_ISL_676 | ···················· | ···················· | ···················· | ···················· | ···················· | ···················· |
| U | 4567 | hCoV-19/USA/NY-Wadsworth-29254-01/2020\|EPI_ISL_676 | ···················· | ···················· | ···················· | ···················· | ···················· | ···················· |
| U | 4568 | hCoV-19/USA/NY-Wadsworth-30118-01/2020\|EPI_ISL_676 | ···················· | ···················· | ···················· | ···················· | ···················· | ···················· |
| U | 4569 | hCoV-19/USA/NY-Wadsworth-30234-01/2020\|EPI_ISL_676 | ···················· | ···················· | ···················· | ···················· | ···················· | ···················· |
| U | 4570 | hCoV-19/USA/NY-Wadsworth-30331-01/2020\|EPI_ISL_676 | ···················· | ···················· | ···················· | ···················· | ···················· | ···················· |
| U | 4571 | hCoV-19/USA/NY-Wadsworth-30491-01/2020\|EPI_ISL_676 | ···················· | ···················· | ···················· | ···················· | ···················· | ···················· |
| U | 4572 | hCoV-19/USA/NY-Wadsworth-30514-01/2020\|EPI_ISL_676 | ···················· | ···················· | ···················· | ···················· | ···················· | ···················· |
| U | 4573 | hCoV-19/USA/NY-Wadsworth-30522-01/2020\|EPI_ISL_676 | ···················· | ···················· | ···················· | ···················· | ···················· | ···················· |
| U | 4574 | hCoV-19/USA/NY-Wadsworth-30605-01/2020\|EPI_ISL_676 | ···················· | ···················· | ···················· | ···················· | ···················· | ···················· |
| U | 4575 | hCoV-19/USA/NY-Wadsworth-30942-01/2020\|EPI_ISL_676 | ···················· | ···················· | ···················· | ···················· | ···················· | ···················· |
| U | 4576 | hCoV-19/USA/NY-Wadsworth-31105-01/2020\|EPI_ISL_676 | ···················· | ···················· | ···················· | ···················· | ···················· | ···················· |
| U | 4577 | hCoV-19/USA/NY-Wadsworth-31124-01/2020\|EPI_ISL_676 | ···················· | ···················· | ···················· | ···················· | ···················· | ···················· |
| U | 4578 | hCoV-19/USA/NY-Wadsworth-31163-01/2020\|EPI_ISL_676 | ···················· | ···················· | ···················· | ···················· | ···················· | ···················· |
| U | 4579 | hCoV-19/USA/NY-Wadsworth-31741-01/2020\|EPI_ISL_676 | ···················· | ···················· | ···················· | ···················· | ···················· | ···················· |

U 4580 hCoV-19/Saudi Arabia/KAUST-MADINAH1129/2020|EPI_IS ···················· ···················· ···················· ···················· ···················· ····················
U 4582 hCoV-19/Saudi Arabia/KAUST-MADINAH1136/2020|EPI_IS ···················· ···················· ···················· ···················· ···················· ····················
U 4583 hCoV-19/Saudi Arabia/KAUST-MADINAH1139/2020|EPI_IS ···················· ·A·················· ···················· ···················· ···················· ···A················
U 4584 hCoV-19/Saudi Arabia/KAUST-MADINAH1140/2020|EPI_IS ···················· ·A·················· ···················· ···················· ···················· ···A················
U 4585 hCoV-19/Saudi Arabia/KAUST-MADINAH1141/2020|EPI_IS ···················· ·A·················· ···················· ···················· ···················· ···A················
U 4586 hCoV-19/Saudi Arabia/KAUST-MADINAH1142/2020|EPI_IS ···················· ·A·················· ···················· ···················· ···················· ···A················
U 4587 hCoV-19/Saudi Arabia/KAUST-MADINAH1143/2020|EPI_IS ···················· ·A·················· ···················· ···················· ···················· ···A················
U 4588 hCoV-19/Saudi Arabia/KAUST-MADINAH1144/2020|EPI_IS ···················· ·A·················· ···················· ···················· ···················· ···A················
U 4589 hCoV-19/Saudi Arabia/KAUST-MADINAH1145/2020|EPI_IS ···················· ·A·················· ···················· ···················· ···················· ···A················
U 4590 hCoV-19/Saudi Arabia/KAUST-MADINAH1146/2020|EPI_IS ···················· ·A·················· ···················· ···················· ···················· ···A················
U 4591 hCoV-19/Saudi Arabia/KAUST-MADINAH1147/2020|EPI_IS ···················· ·A·················· ···················· ···················· ···················· ···A················
U 4592 hCoV-19/Wales/PHWC-48FBE6/2020|EPI_ISL_681201|2020 ···················· ···················· ···················· ···················· ···················· ····················
U 4593 hCoV-19/Wales/PHWC-48FBF5/2020|EPI_ISL_681202|2020 ···················· ···················· ···················· ···················· ···················· ····················
U 4594 hCoV-19/Wales/PHWC-48FC10/2020|EPI_ISL_681203|2020 ···················· ···················· ···················· ···················· ···················· ····················
U 4598 hCoV-19/Wales/PHWC-48FC6B/2020|EPI_ISL_681208|2020 ···················· ···················· ···················· ···················· ···················· ····················
U 4599 hCoV-19/Bahrain/920455954_S8_L001/2020|EPI_ISL_681 ···················· ···················· ···················· ···················· ···················· ····················
U 4600 hCoV-19/Bahrain/920455640_S10_L001/2020|EPI_ISL_68 ···················· ···················· ···················· ···················· ···················· ····················
U 4601 hCoV-19/Denmark/DCGC-12349/2020|EPI_ISL_682530|202 ···················· ···················· ···················· ···················· ···················· ····················
U 4602 hCoV-19/Denmark/DCGC-12350/2020|EPI_ISL_682531|202 ···················· ·A·················· ···················· ···················· ···················· ····················
U 4603 hCoV-19/Denmark/DCGC-12351/2020|EPI_ISL_682532|202 ···················· ···················· ···················· ···················· ···················· ····················
U 4604 hCoV-19/Denmark/DCGC-12352/2020|EPI_ISL_682533|202 ···················· ···················· ···················· ···················· ···················· ····················
U 4605 hCoV-19/Denmark/DCGC-12353/2020|EPI_ISL_682534|202 ···················· ···················· ···················· ···················· ···················· ····················
U 4606 hCoV-19/Denmark/DCGC-12354/2020|EPI_ISL_682535|202 ···················· ···················· ···················· ···················· ···················· ····················
U 4607 hCoV-19/Denmark/DCGC-12356/2020|EPI_ISL_682537|202 ···················· ·A·················· ···················· ···················· ···················· ····················
U 4608 hCoV-19/Denmark/DCGC-12358/2020|EPI_ISL_682539|202 ···················· ···················· ···················· ···················· ···················· ····················
U 4609 hCoV-19/England/MILK-B85E95/2020|EPI_ISL_674000|20 ···················· ···················· ···················· ···················· ···················· ····················
U 4610 hCoV-19/England/MILK-B86447/2020|EPI_ISL_674001|20 ···················· ·A·················· ···················· ···················· ···················· ···A················
U 4611 hCoV-19/England/MILK-B8D46E/2020|EPI_ISL_674080|20 ···················· ···················· ···················· ···················· ···················· ····················
U 4612 hCoV-19/England/MILK-B85D7A/2020|EPI_ISL_674081|20 ···················· ···················· ···················· ···················· ···················· ····················
U 4613 hCoV-19/England/MILK-B8E61C/2020|EPI_ISL_674085|20 ···················· ···················· ···················· ···················· ···················· ····················
U 4614 hCoV-19/England/MILK-B8D52F/2020|EPI_ISL_674088|20 ···················· ···················· ···················· ···················· ···················· ····················
U 4615 hCoV-19/England/MILK-B85EFF/2020|EPI_ISL_674089|20 ···················· ···················· ···················· ···················· ···················· ····················
U 4616 hCoV-19/England/MILK-B85D89/2020|EPI_ISL_674090|20 ···················· ···················· ···················· ···················· ···················· ····················
U 4617 hCoV-19/England/MILK-B8D20A/2020|EPI_ISL_674091|20 ···················· ···················· ···················· ···················· ···················· ····················
U 4618 hCoV-19/England/MILK-B8D361/2020|EPI_ISL_674093|20 ···················· ···················· ···················· ···················· ···················· ····················
U 4619 hCoV-19/England/MILK-B86456/2020|EPI_ISL_674094|20 ···················· ···················· ···················· ···················· ···················· ····················
U 4620 hCoV-19/England/MILK-B8E52E/2020|EPI_ISL_674095|20 ···················· ···················· ···················· ···················· ···················· ····················
U 4621 hCoV-19/England/MILK-B86131/2020|EPI_ISL_674096|20 ···················· ···················· ···················· ···················· ···················· ····················
U 4622 hCoV-19/England/MILK-B86966/2020|EPI_ISL_674097|20 ···················· ···················· ···················· ···················· ···················· ····················
U 4623 hCoV-19/England/MILK-B8D228/2020|EPI_ISL_674098|20 ···················· ···················· ···················· ···················· ···················· ····················
U 4624 hCoV-19/England/MILK-B8D53E/2020|EPI_ISL_674099|20 ···················· ···················· ···················· ···················· ···················· ····················
U 4625 hCoV-19/England/ALDP-B82F3B/2020|EPI_ISL_675314|20 ···················· ···················· ···················· ···················· ···················· ····················
U 4626 hCoV-19/England/ALDP-B83C9D/2020|EPI_ISL_675315|20 ···················· ···················· ···················· ···················· ···················· ····················
U 4627 hCoV-19/England/ALDP-B84643/2020|EPI_ISL_675316|20 ···················· ···················· ···················· ···················· ···················· ····················
U 4628 hCoV-19/England/MILK-B8F1FC/2020|EPI_ISL_675317|20 ···················· ···················· ···················· ···················· ···················· ····················
U 4629 hCoV-19/England/MILK-B8F0C2/2020|EPI_ISL_675318|20 ···················· ···················· ···················· ···················· ···················· ····················
U 4630 hCoV-19/England/MILK-B8F2BD/2020|EPI_ISL_675320|20 ···················· ···················· ···················· ···················· ···················· ····················
U 4631 hCoV-19/England/ALDP-B8378D/2020|EPI_ISL_675321|20 ···················· ···················· ···················· ···················· ···················· ····················
U 4632 hCoV-19/England/ALDP-B837D8/2020|EPI_ISL_675322|20 ···················· ···················· ···················· ···················· ···················· ····················
U 4633 hCoV-19/England/ALDP-B82CCB/2020|EPI_ISL_675323|20 ···················· ···················· ···················· ···················· ·T·················· ····················
U 4634 hCoV-19/England/ALDP-B83750/2020|EPI_ISL_675324|20 ···················· ···················· ···················· ···················· ······T············· ····················
U 4635 hCoV-19/England/ALDP-B842D6/2020|EPI_ISL_675328|20 ···················· ·A·················· ···················· ···················· ···················· ···A················
U 4636 hCoV-19/USA/TX-DSHS-1480/2020|EPI_ISL_676645|2020- ···················· ·A·················· ···················· ···················· ···················· ····················
U 4637 hCoV-19/USA/TX-DSHS-1484/2020|EPI_ISL_676646|2020- ···················· ·A·················· ···················· ···················· ···················· ····················
U 4638 hCoV-19/USA/NY-Wadsworth-15538-01/2020|EPI_ISL_676 ···················· ···················· ···················· ···················· ···················· ····················
U 4639 hCoV-19/USA/NY-Wadsworth-45587-01/2020|EPI_ISL_676 ···················· ···················· ···················· ···················· ···················· ····················
U 4640 hCoV-19/USA/NY-Wadsworth-42205-01/2020|EPI_ISL_676 ···················· ···················· ···················· ···················· ···················· ····················
U 4641 hCoV-19/Saudi Arabia/KAUST-MADINAH1117/2020|EPI_IS ···················· ···················· ···················· ···················· ···················· ····················
U 4642 hCoV-19/Saudi Arabia/KAUST-MADINAH1118/2020|EPI_IS ···················· ·A·················· ···················· ···················· ···················· ···A················
U 4643 hCoV-19/Saudi Arabia/KAUST-MADINAH1119/2020|EPI_IS ···················· ·A·················· ···················· ···················· ···················· ···A················
U 4644 hCoV-19/Saudi Arabia/KAUST-MADINAH1120/2020|EPI_IS ···················· ···················· ···················· ···················· ···················· ····················
U 4645 hCoV-19/Saudi Arabia/KAUST-MADINAH1121/2020|EPI_IS ···················· ·A·················· ···················· ···················· ···················· ···A················
U 4646 hCoV-19/Saudi Arabia/KAUST-MADINAH1122/2020|EPI_IS ···················· ·A·················· ···················· ···················· ···················· ···A················
U 4647 hCoV-19/Saudi Arabia/KAUST-MADINAH1123/2020|EPI_IS ···················· ·A·················· ···················· ···················· ···················· ···A················
U 4648 hCoV-19/Saudi Arabia/KAUST-MADINAH1124/2020|EPI_IS ···················· ·A·················· ···················· ···················· ···················· ···A················
U 4649 hCoV-19/Saudi Arabia/KAUST-MADINAH1126/2020|EPI_IS ···················· ···················· ···················· ···················· ···················· ···A················
U 4650 hCoV-19/Saudi Arabia/KAUST-MADINAH1127/2020|EPI_IS ···················· ···················· ···················· ···················· ···················· ····················
U 4651 hCoV-19/Saudi Arabia/KAUST-MADINAH1131/2020|EPI_IS ···················· ·A·················· ···················· ···················· ···················· ···A················
U 4652 hCoV-19/Wales/PHWC-48F4BE/2020|EPI_ISL_681100|2020 ···················· ···················· ···················· ···················· ···················· ····················
U 4653 hCoV-19/Wales/PHWC-48F4CD/2020|EPI_ISL_681101|2020 ···················· ···················· ···················· ···················· ···················· ····················
U 4654 hCoV-19/Wales/PHWC-48F4DC/2020|EPI_ISL_681102|2020 ···················· ···················· ···················· ···················· ···················· ····················
U 4655 hCoV-19/Wales/PHWC-48F4FA/2020|EPI_ISL_681103|2020 ···················· ···················· ···················· ···················· ···················· ····················
U 4656 hCoV-19/Wales/PHWC-48F506/2020|EPI_ISL_681104|2020 ···················· ···················· ···················· ···················· ···················· ····················
U 4657 hCoV-19/Wales/PHWC-48F515/2020|EPI_ISL_681105|2020 ···················· ···················· ···················· ···················· ···················· ····················
U 4658 hCoV-19/Wales/PHWC-48F533/2020|EPI_ISL_681107|2020 ···················· ···················· ···················· ···················· ···················· ····················
U 4660 hCoV-19/Wales/PHWC-48F58E/2020|EPI_ISL_681111|2020 ···················· ···················· ···················· ···················· ···················· ····················
U 4661 hCoV-19/Wales/PHWC-48F5AC/2020|EPI_ISL_681112|2020 ···················· ···················· ···················· ···················· ···················· ····················
U 4662 hCoV-19/Wales/PHWC-48F5D9/2020|EPI_ISL_681114|2020 ···················· ···················· ···················· ···················· ···················· ····················
U 4665 hCoV-19/Wales/PHWC-48F603/2020|EPI_ISL_681117|2020 ···················· ···················· ···················· ···················· ···················· ····················
U 4666 hCoV-19/Wales/PHWC-48F630/2020|EPI_ISL_681118|2020 ···················· ···················· ···················· ···················· ···················· ····················

| | | | | | | | | |
|---|---|---|---|---|---|---|---|---|
| U | 4667 | hCoV-19/Wales/PHWC-48F66D/2020\|EPI_ISL_681120\|2020 | ···················· | ···················· | ···················· | ···················· | ···················· | ···················· |
| U | 4668 | hCoV-19/Wales/PHWC-48F67C/2020\|EPI_ISL_681121\|2020 | ···················· | ···················· | ···················· | ···················· | ··A················· | ···················· |
| U | 4669 | hCoV-19/Wales/PHWC-48F68B/2020\|EPI_ISL_681122\|2020 | ···················· | ···················· | ···················· | ···················· | ···················· | ···················· |
| U | 4670 | hCoV-19/Wales/PHWC-48F69A/2020\|EPI_ISL_681123\|2020 | ···················· | ···················· | ···················· | ···················· | ···················· | ···················· |
| U | 4671 | hCoV-19/Wales/PHWC-48F6C7/2020\|EPI_ISL_681126\|2020 | ···················· | ···················· | ···················· | ···················· | ···················· | ···················· |
| U | 4672 | hCoV-19/Wales/PHWC-48F6D6/2020\|EPI_ISL_681127\|2020 | ···················· | ···················· | ···················· | ···················· | ···················· | ···················· |
| U | 4673 | hCoV-19/Wales/PHWC-48F6E5/2020\|EPI_ISL_681128\|2020 | ···················· | ···················· | ···················· | ···················· | ···················· | ···················· |
| U | 4674 | hCoV-19/Wales/PHWC-48F6F4/2020\|EPI_ISL_681129\|2020 | ···················· | ···················· | ···················· | ···················· | ···················· | ···················· |
| U | 4675 | hCoV-19/Wales/PHWC-48F700/2020\|EPI_ISL_681130\|2020 | ···················· | ···················· | ···················· | ···················· | ···················· | ···················· |
| U | 4676 | hCoV-19/Wales/PHWC-48F73D/2020\|EPI_ISL_681132\|2020 | ···················· | ···················· | ···················· | ···················· | ···················· | ···················· |
| U | 4677 | hCoV-19/Wales/PHWC-48F75B/2020\|EPI_ISL_681133\|2020 | ···················· | ···················· | ···················· | ···················· | ···················· | ···················· |
| U | 4678 | hCoV-19/Wales/PHWC-48F76A/2020\|EPI_ISL_681134\|2020 | ···················· | ···················· | ···················· | ···················· | ···················· | ···················· |
| U | 4679 | hCoV-19/Wales/PHWC-48F788/2020\|EPI_ISL_681135\|2020 | ···················· | ···················· | ···················· | ···················· | ···················· | ···················· |
| U | 4680 | hCoV-19/Wales/PHWC-48F797/2020\|EPI_ISL_681136\|2020 | ···················· | ···················· | ···················· | ···················· | ··R················· | ···················· |
| U | 4681 | hCoV-19/Wales/PHWC-48F7A6/2020\|EPI_ISL_681137\|2020 | ···················· | ···················· | ···················· | ···················· | ···················· | ···················· |
| U | 4683 | hCoV-19/Wales/PHWC-48F7D3/2020\|EPI_ISL_681140\|2020 | ···················· | ···················· | ···················· | ···················· | ···················· | ···················· |
| U | 4684 | hCoV-19/Wales/PHWC-48F7F1/2020\|EPI_ISL_681142\|2020 | ···················· | ···················· | ···················· | ···················· | ···················· | ···················· |
| U | 4685 | hCoV-19/Wales/PHWC-48F81C/2020\|EPI_ISL_681144\|2020 | ···················· | ···················· | ···················· | ···················· | ···················· | ···················· |
| U | 4687 | hCoV-19/Wales/PHWC-48F867/2020\|EPI_ISL_681148\|2020 | ···················· | ···················· | ···················· | ···················· | ···················· | ···················· |
| U | 4688 | hCoV-19/Wales/PHWC-48F876/2020\|EPI_ISL_681149\|2020 | ···················· | ···················· | ···················· | ···················· | ···················· | ···················· |
| U | 4689 | hCoV-19/Wales/PHWC-48F885/2020\|EPI_ISL_681150\|2020 | ···················· | ···················· | ···················· | ···················· | ···················· | ···················· |
| U | 4690 | hCoV-19/Wales/PHWC-48F894/2020\|EPI_ISL_681151\|2020 | ···················· | ···················· | ···················· | ···················· | ···················· | ···················· |
| U | 4691 | hCoV-19/Wales/PHWC-48F8A3/2020\|EPI_ISL_681152\|2020 | ···················· | ···················· | ···················· | ···················· | ···················· | ···················· |
| U | 4692 | hCoV-19/Wales/PHWC-48F8B2/2020\|EPI_ISL_681153\|2020 | ···················· | ···················· | ···················· | ···················· | ···················· | ···················· |
| U | 4693 | hCoV-19/Wales/PHWC-48F8C1/2020\|EPI_ISL_681154\|2020 | ···················· | ···················· | ···················· | ···················· | ···················· | ···················· |
| U | 4694 | hCoV-19/Wales/PHWC-48F8EF/2020\|EPI_ISL_681156\|2020 | ···················· | ···················· | ···················· | ···················· | ···················· | ···················· |
| U | 4695 | hCoV-19/Wales/PHWC-48F8FE/2020\|EPI_ISL_681157\|2020 | ···················· | ···················· | ···················· | ···················· | ···················· | ···················· |
| U | 4696 | hCoV-19/Wales/PHWC-48F90A/2020\|EPI_ISL_681158\|2020 | ···················· | ···················· | ···················· | ···················· | ···················· | ···················· |
| U | 4697 | hCoV-19/Wales/PHWC-48F919/2020\|EPI_ISL_681159\|2020 | ···················· | ···················· | ···················· | ···················· | ···················· | ···················· |
| U | 4698 | hCoV-19/Wales/PHWC-48F937/2020\|EPI_ISL_681161\|2020 | ···················· | ···················· | ···················· | ···················· | ···················· | ···················· |
| U | 4699 | hCoV-19/Wales/PHWC-48F955/2020\|EPI_ISL_681163\|2020 | ···················· | ···················· | ···················· | ···················· | ···················· | ···················· |
| U | 4700 | hCoV-19/Wales/PHWC-48F964/2020\|EPI_ISL_681164\|2020 | ···················· | ···················· | ···················· | ···················· | ···················· | ···················· |
| U | 4701 | hCoV-19/Wales/PHWC-48F973/2020\|EPI_ISL_681165\|2020 | ···················· | ···················· | ···················· | ···················· | ···················· | ···················· |
| U | 4702 | hCoV-19/Denmark/DCGC-12229/2020\|EPI_ISL_682410\|202 | ···················· | ···················· | ···················· | ···················· | ···················· | ···················· |
| U | 4703 | hCoV-19/Denmark/DCGC-12231/2020\|EPI_ISL_682412\|202 | ···················· | ···················· | ···················· | ···················· | ···················· | ···················· |
| U | 4704 | hCoV-19/Denmark/DCGC-12232/2020\|EPI_ISL_682413\|202 | ···················· | ···················· | ···················· | ···················· | ···················· | ···················· |
| U | 4705 | hCoV-19/Denmark/DCGC-12235/2020\|EPI_ISL_682416\|202 | ···················· | ···················· | ···················· | ···················· | ···················· | ···················· |
| U | 4706 | hCoV-19/Denmark/DCGC-12237/2020\|EPI_ISL_682418\|202 | ···················· | ···················· | ···················· | ···················· | ···················· | ···················· |
| U | 4707 | hCoV-19/Denmark/DCGC-12239/2020\|EPI_ISL_682420\|202 | ···················· | ···················· | ···················· | ···················· | ···················· | ···················· |
| U | 4708 | hCoV-19/Denmark/DCGC-12240/2020\|EPI_ISL_682421\|202 | ···················· | ···················· | ···················· | ···················· | ···················· | ···················· |
| U | 4709 | hCoV-19/Denmark/DCGC-12241/2020\|EPI_ISL_682422\|202 | ···················· | ···················· | ···················· | ···················· | ···················· | ···················· |
| U | 4710 | hCoV-19/Denmark/DCGC-12242/2020\|EPI_ISL_682423\|202 | ···················· | ···················· | ···················· | ···················· | ···················· | ···················· |
| U | 4711 | hCoV-19/Denmark/DCGC-12243/2020\|EPI_ISL_682424\|202 | ···················· | ···················· | ···················· | ···················· | ···················· | ···················· |
| U | 4712 | hCoV-19/Denmark/DCGC-12244/2020\|EPI_ISL_682425\|202 | ···················· | ···················· | ···················· | ···················· | ···················· | ···················· |
| U | 4713 | hCoV-19/Denmark/DCGC-12246/2020\|EPI_ISL_682427\|202 | ···················· | ···················· | ···················· | ···················· | ···················· | ···················· |
| U | 4714 | hCoV-19/Denmark/DCGC-12247/2020\|EPI_ISL_682428\|202 | ···················· | ·A·················· | ···················· | ···················· | ···················· | ···················· |
| U | 4715 | hCoV-19/Denmark/DCGC-12248/2020\|EPI_ISL_682429\|202 | ···················· | ···················· | ···················· | ···················· | ···················· | ···················· |
| U | 4716 | hCoV-19/Denmark/DCGC-12250/2020\|EPI_ISL_682431\|202 | ···················· | ···················· | ···················· | ···················· | ···················· | ···················· |
| U | 4717 | hCoV-19/Denmark/DCGC-12251/2020\|EPI_ISL_682432\|202 | ···················· | ···················· | ···················· | ···················· | ···················· | ···················· |
| U | 4718 | hCoV-19/Denmark/DCGC-12252/2020\|EPI_ISL_682433\|202 | ···················· | ···················· | ···················· | ···················· | ···················· | ···················· |
| U | 4719 | hCoV-19/Denmark/DCGC-12253/2020\|EPI_ISL_682434\|202 | ···················· | ···················· | ···················· | ···················· | ···················· | ···················· |
| U | 4720 | hCoV-19/Denmark/DCGC-12254/2020\|EPI_ISL_682435\|202 | ···················· | ···················· | ···················· | ···················· | ···················· | ···················· |
| U | 4721 | hCoV-19/Denmark/DCGC-12255/2020\|EPI_ISL_682436\|202 | ···················· | ···················· | ···················· | ···················· | ···················· | ···················· |
| U | 4722 | hCoV-19/Denmark/DCGC-12256/2020\|EPI_ISL_682437\|202 | ···················· | ···················· | ···················· | ···················· | ···················· | ···················· |
| U | 4723 | hCoV-19/Denmark/DCGC-12257/2020\|EPI_ISL_682438\|202 | ···················· | ···················· | ···················· | ···················· | ···················· | ···················· |
| U | 4724 | hCoV-19/Denmark/DCGC-12259/2020\|EPI_ISL_682440\|202 | ···················· | ···················· | ···················· | ···················· | ···················· | ···················· |
| U | 4725 | hCoV-19/Denmark/DCGC-12260/2020\|EPI_ISL_682441\|202 | ···················· | ···················· | ···················· | ···················· | ···················· | ···················· |
| U | 4726 | hCoV-19/Denmark/DCGC-12264/2020\|EPI_ISL_682445\|202 | ···················· | ···················· | ···················· | ···················· | ···················· | ···················· |
| U | 4727 | hCoV-19/Denmark/DCGC-12265/2020\|EPI_ISL_682446\|202 | ···················· | ···················· | ···················· | ···················· | ···················· | ···················· |
| U | 4728 | hCoV-19/Denmark/DCGC-12266/2020\|EPI_ISL_682447\|202 | ···················· | ···················· | ···················· | ···················· | ···················· | ···················· |
| U | 4729 | hCoV-19/Denmark/DCGC-12267/2020\|EPI_ISL_682448\|202 | ···················· | ···················· | ···················· | ···················· | ···················· | ···················· |
| U | 4730 | hCoV-19/Denmark/DCGC-12268/2020\|EPI_ISL_682449\|202 | ···················· | ···················· | ···················· | ···················· | ···················· | ···················· |
| U | 4731 | hCoV-19/Denmark/DCGC-12269/2020\|EPI_ISL_682450\|202 | ···················· | ···················· | ···················· | ···················· | ···················· | ···················· |
| U | 4732 | hCoV-19/Denmark/DCGC-12270/2020\|EPI_ISL_682451\|202 | ···················· | ···················· | ···················· | ···················· | ···················· | ···················· |
| U | 4733 | hCoV-19/Denmark/DCGC-12271/2020\|EPI_ISL_682452\|202 | ···················· | ·A·················· | ···················· | ···················· | ···················· | ···A················ |
| U | 4734 | hCoV-19/Denmark/DCGC-12274/2020\|EPI_ISL_682455\|202 | ···················· | ···················· | ···················· | ···················· | ···················· | ···················· |
| U | 4735 | hCoV-19/Denmark/DCGC-12275/2020\|EPI_ISL_682456\|202 | ···················· | ···················· | ···················· | ···················· | ···················· | ···················· |
| U | 4736 | hCoV-19/Denmark/DCGC-12276/2020\|EPI_ISL_682457\|202 | ···················· | ···················· | ···················· | ···················· | ···················· | ···················· |
| U | 4737 | hCoV-19/Denmark/DCGC-12277/2020\|EPI_ISL_682458\|202 | ···················· | ···················· | ···················· | ···················· | ···················· | ···················· |
| U | 4738 | hCoV-19/Denmark/DCGC-12278/2020\|EPI_ISL_682459\|202 | ···················· | ···················· | ···················· | ···················· | ···················· | ···················· |
| U | 4739 | hCoV-19/Denmark/DCGC-12279/2020\|EPI_ISL_682460\|202 | ···················· | ···A················ | ···················· | ···················· | ···················· | ···················· |
| U | 4740 | hCoV-19/Denmark/DCGC-12280/2020\|EPI_ISL_682461\|202 | ···················· | ···················· | ···················· | ···················· | ···················· | ···················· |
| U | 4741 | hCoV-19/Denmark/DCGC-12281/2020\|EPI_ISL_682462\|202 | ···················· | ···················· | ···················· | ···················· | ···················· | ···················· |
| U | 4742 | hCoV-19/Denmark/DCGC-12282/2020\|EPI_ISL_682463\|202 | ···················· | ···················· | ···················· | ···················· | ···················· | ···················· |
| U | 4743 | hCoV-19/Denmark/DCGC-12283/2020\|EPI_ISL_682464\|202 | ···················· | ···················· | ···················· | ···················· | ···················· | ···················· |
| U | 4744 | hCoV-19/Denmark/DCGC-12287/2020\|EPI_ISL_682468\|202 | ···················· | ···················· | ···················· | ···················· | ···················· | ···················· |
| U | 4745 | hCoV-19/Denmark/DCGC-12289/2020\|EPI_ISL_682470\|202 | ···················· | ···················· | ···················· | ···················· | ···················· | ···················· |
| U | 4746 | hCoV-19/Denmark/DCGC-12291/2020\|EPI_ISL_682472\|202 | ···················· | ···················· | ···················· | ···················· | ···················· | ···················· |
| U | 4747 | hCoV-19/Denmark/DCGC-12293/2020\|EPI_ISL_682474\|202 | ···················· | ···················· | ···················· | ···················· | ···················· | ···················· |
| U | 4748 | hCoV-19/Denmark/DCGC-12294/2020\|EPI_ISL_682475\|202 | ···················· | ···················· | ···················· | ···················· | ···················· | ···················· |

| | | | | | | | |
|---|---|---|---|---|---|---|---|
| U | 4749 hCoV-19/Denmark/DCGC-12295/2020\|EPI_ISL_682476\|202 | ···················· | ···················· | ···················· | ···················· | ···················· | ···················· |
| U | 4750 hCoV-19/Denmark/DCGC-12296/2020\|EPI_ISL_682477\|202 | ···················· | ···················· | ···················· | ···················· | ···················· | ···················· |
| U | 4751 hCoV-19/Denmark/DCGC-12297/2020\|EPI_ISL_682478\|202 | ···················· | ···················· | ···················· | ···················· | ···················· | ···················· |
| U | 4752 hCoV-19/Denmark/DCGC-12298/2020\|EPI_ISL_682479\|202 | ···················· | ···················· | ···················· | ···················· | ···················· | ···················· |
| U | 4753 hCoV-19/Denmark/DCGC-12300/2020\|EPI_ISL_682481\|202 | ···················· | ···················· | ···················· | ···················· | ···················· | ···················· |
| U | 4754 hCoV-19/Denmark/DCGC-12301/2020\|EPI_ISL_682482\|202 | ···················· | ···················· | ···················· | ···················· | ···················· | ···················· |
| U | 4755 hCoV-19/Denmark/DCGC-12305/2020\|EPI_ISL_682486\|202 | ···················· | ···················· | ···················· | ···················· | ···················· | ···················· |
| U | 4756 hCoV-19/Denmark/DCGC-12306/2020\|EPI_ISL_682487\|202 | ···················· | ···················· | ···················· | ···················· | ···················· | ···················· |
| U | 4757 hCoV-19/Denmark/DCGC-12307/2020\|EPI_ISL_682488\|202 | ···················· | ···················· | ···················· | ···················· | ···················· | ···················· |
| U | 4758 hCoV-19/Denmark/DCGC-12308/2020\|EPI_ISL_682489\|202 | ···················· | ···················· | ···················· | ···················· | ···················· | ···················· |
| U | 4759 hCoV-19/Denmark/DCGC-12310/2020\|EPI_ISL_682491\|202 | ···················· | ···················· | ···················· | ···················· | ···················· | ···················· |
| U | 4760 hCoV-19/Denmark/DCGC-12311/2020\|EPI_ISL_682492\|202 | ···················· | ···················· | ···················· | ···················· | ·····T·············· | ···················· |
| U | 4761 hCoV-19/Denmark/DCGC-12313/2020\|EPI_ISL_682494\|202 | ···················· | ·A·················· | ···················· | ···················· | ···················· | ··A················· |
| U | 4762 hCoV-19/Denmark/DCGC-12314/2020\|EPI_ISL_682495\|202 | ···················· | ···················· | ···················· | ···················· | ···················· | ···················· |
| U | 4763 hCoV-19/Denmark/DCGC-12315/2020\|EPI_ISL_682496\|202 | ···················· | ···················· | ···················· | ···················· | ···················· | ···················· |
| U | 4764 hCoV-19/Denmark/DCGC-12319/2020\|EPI_ISL_682500\|202 | ···················· | ···················· | ···················· | ···················· | ···················· | ···················· |
| U | 4765 hCoV-19/Denmark/DCGC-12320/2020\|EPI_ISL_682501\|202 | ···················· | ···················· | ···················· | ···················· | ···················· | ···················· |
| U | 4766 hCoV-19/Denmark/DCGC-12321/2020\|EPI_ISL_682502\|202 | ···················· | ···················· | ···················· | ···················· | ···················· | ···················· |
| U | 4767 hCoV-19/Denmark/DCGC-12322/2020\|EPI_ISL_682503\|202 | ···················· | ···················· | ···················· | ···················· | ···················· | ···················· |
| U | 4768 hCoV-19/Denmark/DCGC-12323/2020\|EPI_ISL_682504\|202 | ···················· | ···················· | ···················· | ···················· | ···················· | ···················· |
| U | 4769 hCoV-19/Denmark/DCGC-12324/2020\|EPI_ISL_682505\|202 | ···················· | ···················· | ···················· | ···················· | ···················· | ···················· |
| U | 4770 hCoV-19/Denmark/DCGC-12329/2020\|EPI_ISL_682510\|202 | ···················· | ···················· | ···················· | ···················· | ···················· | ···················· |
| U | 4771 hCoV-19/Denmark/DCGC-12331/2020\|EPI_ISL_682512\|202 | ···················· | ···················· | ···················· | ···················· | ···················· | ···················· |
| U | 4772 hCoV-19/Denmark/DCGC-12333/2020\|EPI_ISL_682514\|202 | ···················· | ···················· | ···················· | ···················· | ···················· | ···················· |
| U | 4773 hCoV-19/Denmark/DCGC-12334/2020\|EPI_ISL_682515\|202 | ···················· | ···················· | ···················· | ···················· | ···················· | ···················· |
| U | 4774 hCoV-19/Denmark/DCGC-12335/2020\|EPI_ISL_682516\|202 | ···················· | ···················· | ···················· | ···················· | ···················· | ···················· |
| U | 4775 hCoV-19/Denmark/DCGC-12336/2020\|EPI_ISL_682517\|202 | ···················· | ···················· | ···················· | ···················· | ···················· | ···················· |
| U | 4776 hCoV-19/Denmark/DCGC-12337/2020\|EPI_ISL_682518\|202 | ···················· | ···················· | ···················· | ···················· | ···················· | ···················· |
| U | 4777 hCoV-19/Denmark/DCGC-12338/2020\|EPI_ISL_682519\|202 | ···················· | ···················· | ···················· | ···················· | ···················· | ···················· |
| U | 4778 hCoV-19/Denmark/DCGC-12339/2020\|EPI_ISL_682520\|202 | ···················· | ·A·················· | ···················· | ···················· | ···················· | ···················· |
| U | 4779 hCoV-19/Denmark/DCGC-12341/2020\|EPI_ISL_682522\|202 | ···················· | ···················· | ···················· | ···················· | ···················· | ···················· |
| U | 4780 hCoV-19/Denmark/DCGC-12342/2020\|EPI_ISL_682523\|202 | ···················· | ···················· | ···················· | ···················· | ···················· | ···················· |
| U | 4781 hCoV-19/Denmark/DCGC-12343/2020\|EPI_ISL_682524\|202 | ···················· | ···················· | ···················· | ···················· | ···················· | ···················· |
| U | 4782 hCoV-19/Denmark/DCGC-12344/2020\|EPI_ISL_682525\|202 | ···················· | ···················· | ···················· | ···················· | ···················· | ···················· |
| U | 4783 hCoV-19/Denmark/DCGC-12345/2020\|EPI_ISL_682526\|202 | ···················· | ···················· | ···················· | ···················· | ···················· | ···················· |
| U | 4784 hCoV-19/Denmark/DCGC-12346/2020\|EPI_ISL_682527\|202 | ···················· | ···················· | ···················· | ···················· | ···················· | ···················· |
| U | 4785 hCoV-19/Denmark/DCGC-12347/2020\|EPI_ISL_682528\|202 | ···················· | ···················· | ···················· | ···················· | ···················· | ···················· |
| U | 4786 hCoV-19/Denmark/DCGC-12348/2020\|EPI_ISL_682529\|202 | ···················· | ···················· | ···················· | ···················· | ···················· | ···················· |
| U | 4787 hCoV-19/USA/MN-MDH-2111/2020\|EPI_ISL_683740\|2020-1 | ···················· | ···················· | ···················· | ···················· | ···················· | ···················· |
| U | 4788 hCoV-19/USA/MN-MDH-2112/2020\|EPI_ISL_683741\|2020-1 | ···················· | ···················· | ···················· | ···················· | ···················· | ···················· |
| U | 4789 hCoV-19/USA/MN-MDH-2113/2020\|EPI_ISL_683742\|2020-1 | ···················· | ···················· | ···················· | ···················· | ···················· | ···················· |
| U | 4790 hCoV-19/USA/MN-MDH-2114/2020\|EPI_ISL_683743\|2020-1 | ···················· | ···················· | ···················· | ···················· | ···················· | ···················· |
| U | 4791 hCoV-19/USA/MN-MDH-2115/2020\|EPI_ISL_683744\|2020-1 | ···················· | ···················· | ···················· | ···················· | ···················· | ···················· |
| U | 4792 hCoV-19/USA/MN-MDH-2116/2020\|EPI_ISL_683745\|2020-1 | ···················· | ···················· | ···················· | ···················· | ···················· | ···················· |
| U | 4793 hCoV-19/USA/MN-MDH-2117/2020\|EPI_ISL_683746\|2020-1 | ···················· | ···················· | ···················· | ···················· | ···················· | ···················· |
| U | 4794 hCoV-19/USA/MN-MDH-2118/2020\|EPI_ISL_683747\|2020-1 | ···················· | ···················· | ···················· | ···················· | ···················· | ···················· |
| U | 4795 hCoV-19/USA/MN-MDH-2119/2020\|EPI_ISL_683748\|2020-1 | ···················· | ···················· | ···················· | ···················· | ···················· | ···················· |
| U | 4796 hCoV-19/USA/MN-MDH-2120/2020\|EPI_ISL_683749\|2020-1 | ···················· | ···················· | ···················· | ···················· | ···················· | ···················· |
| U | 4797 hCoV-19/USA/MN-MDH-2121/2020\|EPI_ISL_683750\|2020-1 | ···················· | ···················· | ···················· | ···················· | ···················· | ···················· |
| U | 4798 hCoV-19/USA/NY-NYCPHL-001428/2020\|EPI_ISL_683765\|2 | ···················· | ·A·················· | ···················· | ···················· | ···················· | ···················· |
| U | 4799 hCoV-19/USA/NY-NYCPHL-001455/2020\|EPI_ISL_683766\|2 | ···················· | ···················· | ···················· | ···················· | ···················· | ···················· |
| U | 4800 hCoV-19/USA/NY-NYCPHL-001190/2020\|EPI_ISL_683771\|2 | ···················· | ···················· | ···················· | ···················· | ···················· | ···················· |
| U | 4801 hCoV-19/USA/NY-NYCPHL-001197/2020\|EPI_ISL_683772\|2 | ···················· | ···················· | ···················· | ···················· | ···················· | ···················· |
| U | 4802 hCoV-19/USA/NY-NYCPHL-001198/2020\|EPI_ISL_683773\|2 | ···················· | ·A·················· | ···················· | ···················· | ···················· | ···················· |
| U | 4803 hCoV-19/USA/NY-NYCPHL-001199/2020\|EPI_ISL_683774\|2 | ···················· | ···················· | ···················· | ···················· | ···················· | ···················· |
| U | 4804 hCoV-19/USA/NY-NYCPHL-001200/2020\|EPI_ISL_683775\|2 | ···················· | ···················· | ···················· | ···················· | ···················· | ···················· |
| U | 4805 hCoV-19/USA/NY-NYCPHL-001201/2020\|EPI_ISL_683776\|2 | ···················· | ···················· | ···················· | ···················· | ···················· | ···················· |
| U | 4806 hCoV-19/USA/NY-NYCPHL-001203/2020\|EPI_ISL_683777\|2 | ···················· | ···················· | ···················· | ···················· | ···················· | ···················· |
| U | 4807 hCoV-19/USA/NY-NYCPHL-001204/2020\|EPI_ISL_683778\|2 | ···················· | ·A·················· | ···················· | ···················· | ···················· | ···················· |
| U | 4808 hCoV-19/USA/NY-NYCPHL-001400/2020\|EPI_ISL_683779\|2 | ···················· | ···················· | ···················· | ···················· | ···················· | ···················· |
| U | 4809 hCoV-19/USA/NY-NYCPHL-001401/2020\|EPI_ISL_683780\|2 | ···················· | ···················· | ···················· | ···················· | ···················· | ···················· |
| U | 4810 hCoV-19/USA/NY-NYCPHL-001405/2020\|EPI_ISL_683781\|2 | ···················· | ···················· | ···················· | ···················· | ···················· | ···················· |
| U | 4811 hCoV-19/USA/NY-NYCPHL-001406/2020\|EPI_ISL_683782\|2 | ···················· | ···················· | ···················· | ···················· | ···················· | ···················· |
| U | 4812 hCoV-19/USA/NY-NYCPHL-001407/2020\|EPI_ISL_683783\|2 | ···················· | ·A·················· | ···················· | ···················· | ···················· | ···················· |
| U | 4813 hCoV-19/USA/NY-NYCPHL-001408/2020\|EPI_ISL_683784\|2 | ···················· | ···················· | ···················· | ···················· | ···················· | ···················· |
| U | 4814 hCoV-19/USA/NY-NYCPHL-001411/2020\|EPI_ISL_683785\|2 | ···················· | ···················· | ···················· | ···················· | ···················· | ···················· |
| U | 4815 hCoV-19/USA/NY-NYCPHL-001414/2020\|EPI_ISL_683786\|2 | ···················· | ···················· | ···················· | ···················· | ···················· | ···················· |
| U | 4816 hCoV-19/USA/NY-NYCPHL-001415/2020\|EPI_ISL_683787\|2 | ···················· | ·A·················· | ···················· | ···················· | ···················· | ···················· |
| U | 4817 hCoV-19/USA/NY-NYCPHL-001416/2020\|EPI_ISL_683788\|2 | ···················· | ·A·················· | ···················· | ···················· | ···················· | ···················· |
| U | 4818 hCoV-19/USA/NY-NYCPHL-001417/2020\|EPI_ISL_683789\|2 | ···················· | ···················· | ···················· | ···················· | ···················· | ···················· |
| U | 4819 hCoV-19/USA/NY-NYCPHL-001418/2020\|EPI_ISL_683790\|2 | ···················· | ·A·················· | ···················· | ···················· | ···················· | ···················· |
| U | 4820 hCoV-19/USA/NY-NYCPHL-001420/2020\|EPI_ISL_683791\|2 | ···················· | ···················· | ···················· | ···················· | ···················· | ···················· |
| U | 4821 hCoV-19/USA/NY-NYCPHL-001421/2020\|EPI_ISL_683792\|2 | ···················· | ···················· | ···················· | ···················· | ···················· | ···················· |
| U | 4822 hCoV-19/USA/NY-NYCPHL-001423/2020\|EPI_ISL_683793\|2 | ···················· | ···················· | ···················· | ···················· | ···················· | ···················· |
| U | 4823 hCoV-19/USA/NY-NYCPHL-001424/2020\|EPI_ISL_683794\|2 | ···················· | ···················· | ···················· | ···················· | ···················· | ···················· |
| U | 4824 hCoV-19/USA/NY-NYCPHL-001426/2020\|EPI_ISL_683795\|2 | ···················· | ···················· | ···················· | ···················· | ···················· | ···················· |
| U | 4825 hCoV-19/USA/NY-NYCPHL-001427/2020\|EPI_ISL_683796\|2 | ···················· | ·A·················· | ···················· | ···················· | ···················· | ···················· |
| U | 4826 hCoV-19/USA/NY-NYCPHL-001429/2020\|EPI_ISL_683797\|2 | ···················· | ···················· | ···················· | ···················· | ···················· | ···················· |
| U | 4827 hCoV-19/USA/NY-NYCPHL-001430/2020\|EPI_ISL_683798\|2 | ···················· | ·A·················· | ···················· | ···················· | ···················· | ···················· |
| U | 4828 hCoV-19/USA/NY-NYCPHL-001432/2020\|EPI_ISL_683799\|2 | ···················· | ·A·················· | ···················· | ···················· | ···················· | ···················· |

| | | | | | | | | |
|---|---|---|---|---|---|---|---|---|
| U | 4829 | hCoV-19/USA/NY-NYCPHL-001433/2020\|EPI_ISL_683800\|2 | ···················· | ·A·················· | ···················· | ···················· | ···················· | ···················· |
| U | 4830 | hCoV-19/USA/NY-NYCPHL-001434/2020\|EPI_ISL_683801\|2 | ···················· | ···················· | ···········T········ | ···················· | ···················· | ···················· |
| U | 4831 | hCoV-19/USA/NY-NYCPHL-001435/2020\|EPI_ISL_683802\|2 | ···················· | ·············A······ | ···················· | ···················· | ···················· | ···················· |
| U | 4832 | hCoV-19/USA/NY-NYCPHL-001437/2020\|EPI_ISL_683803\|2 | ···················· | ·A·················· | ···················· | ···················· | ···················· | ···················· |
| U | 4833 | hCoV-19/USA/NY-NYCPHL-001439/2020\|EPI_ISL_683804\|2 | ···················· | ·A·················· | ···················· | ···················· | ···················· | ···················· |
| U | 4834 | hCoV-19/USA/NY-NYCPHL-001440/2020\|EPI_ISL_683805\|2 | ···················· | ·A·················· | ···················· | ···················· | ···················· | ···················· |
| U | 4835 | hCoV-19/USA/NY-NYCPHL-001441/2020\|EPI_ISL_683806\|2 | ···················· | ·A·················· | ···················· | ···················· | ···················· | ···················· |
| U | 4836 | hCoV-19/USA/NY-NYCPHL-001442/2020\|EPI_ISL_683807\|2 | ···················· | ···················· | ···················· | ···················· | ···················· | ···················· |
| U | 4837 | hCoV-19/USA/NY-NYCPHL-001444/2020\|EPI_ISL_683808\|2 | ···················· | ···················· | ···A················ | ···················· | ···················· | ···················· |
| U | 4838 | hCoV-19/USA/NY-NYCPHL-001445/2020\|EPI_ISL_683809\|2 | ···················· | ·A·················· | ···················· | ···················· | ···················· | ···················· |
| U | 4839 | hCoV-19/USA/NY-NYCPHL-001446/2020\|EPI_ISL_683810\|2 | ···················· | ···················· | ···················· | ···················· | ···················· | ···················· |
| U | 4840 | hCoV-19/USA/NY-NYCPHL-001447/2020\|EPI_ISL_683811\|2 | ···················· | ···················· | ···················· | ···················· | ···················· | ···················· |
| U | 4841 | hCoV-19/USA/NY-NYCPHL-001448/2020\|EPI_ISL_683812\|2 | ···················· | ···················· | ···················· | ···················· | ···················· | ···················· |
| U | 4842 | hCoV-19/USA/NY-NYCPHL-001449/2020\|EPI_ISL_683813\|2 | ···················· | ···················· | ···················· | ···················· | ···················· | ···················· |
| U | 4843 | hCoV-19/USA/NY-NYCPHL-001450/2020\|EPI_ISL_683814\|2 | ···················· | ···················· | ···················· | ···················· | ···················· | ···················· |
| U | 4844 | hCoV-19/USA/NY-NYCPHL-001452/2020\|EPI_ISL_683815\|2 | ···················· | ···················· | ···················· | ···················· | ···················· | ···················· |
| U | 4845 | hCoV-19/USA/NY-NYCPHL-001453/2020\|EPI_ISL_683816\|2 | ···················· | ·A·················· | ···················· | ···················· | ···················· | ···················· |
| U | 4846 | hCoV-19/USA/NY-NYCPHL-001454/2020\|EPI_ISL_683817\|2 | ···················· | ···················· | ···················· | ···················· | ···················· | ···················· |
| U | 4847 | hCoV-19/USA/NY-NYCPHL-001456/2020\|EPI_ISL_683818\|2 | ···················· | ·A·················· | ···················· | ···················· | ···················· | ···················· |
| U | 4848 | hCoV-19/USA/NY-NYCPHL-001457/2020\|EPI_ISL_683819\|2 | ···················· | ···················· | ···················· | ···················· | ···················· | ···················· |
| U | 4849 | hCoV-19/USA/NY-NYCPHL-001459/2020\|EPI_ISL_683820\|2 | ···················· | ···················· | ···················· | ···················· | ···················· | ···················· |
| U | 4850 | hCoV-19/USA/NY-NYCPHL-001460/2020\|EPI_ISL_683821\|2 | ···················· | ···················· | ···················· | ···················· | ···················· | ···················· |
| U | 4851 | hCoV-19/USA/NY-NYCPHL-001461/2020\|EPI_ISL_683822\|2 | ···················· | ·A·················· | ···················· | ···················· | ···················· | ···················· |
| U | 4852 | hCoV-19/USA/NY-NYCPHL-001462/2020\|EPI_ISL_683823\|2 | ···················· | ···················· | ···················· | ···················· | ···················· | ···················· |
| U | 4853 | hCoV-19/USA/NY-NYCPHL-001463/2020\|EPI_ISL_683824\|2 | ···················· | ·A·················· | ···················· | ···················· | ···················· | ···················· |
| U | 4854 | hCoV-19/USA/NY-NYCPHL-001464/2020\|EPI_ISL_683825\|2 | ···················· | ·A·················· | ···················· | ···················· | ···················· | ···················· |
| U | 4855 | hCoV-19/USA/NY-NYCPHL-001467/2020\|EPI_ISL_683826\|2 | ···················· | ·A·················· | ···················· | ···················· | ···················· | ···················· |
| U | 4856 | hCoV-19/USA/NY-NYCPHL-001468/2020\|EPI_ISL_683827\|2 | ···················· | ·A·················· | ···················· | ···················· | ···················· | ···················· |
| U | 4857 | hCoV-19/USA/NY-NYCPHL-001469/2020\|EPI_ISL_683828\|2 | ···················· | ···················· | ···················· | ···················· | ···················· | ···················· |
| U | 4858 | hCoV-19/USA/NY-NYCPHL-001470/2020\|EPI_ISL_683829\|2 | ···················· | ·········A·········· | ···················· | ···················· | ···················· | ···················· |
| U | 4859 | hCoV-19/USA/NY-NYCPHL-001471/2020\|EPI_ISL_683830\|2 | ···················· | ···················· | ···················· | ···················· | ···················· | ···················· |
| U | 4860 | hCoV-19/USA/NY-NYCPHL-001473/2020\|EPI_ISL_683831\|2 | ···················· | ···················· | ···················· | ···················· | ···················· | ···················· |
| U | 4861 | hCoV-19/USA/NY-NYCPHL-001476/2020\|EPI_ISL_683832\|2 | ···················· | ·A·················· | ···················· | ···················· | ···················· | ···················· |
| U | 4862 | hCoV-19/USA/NY-NYCPHL-001477/2020\|EPI_ISL_683833\|2 | ···················· | ···················· | ···················· | ···················· | ···················· | ···················· |
| U | 4863 | hCoV-19/USA/NY-NYCPHL-001478/2020\|EPI_ISL_683834\|2 | ···················· | ···················· | ···················· | ···················· | ···················· | ···················· |
| U | 4864 | hCoV-19/Mali/M00960/2020\|EPI_ISL_683835\|2020-06-10 | ···················· | ···················· | ···················· | ···················· | ···················· | ···················· |
| U | 4865 | hCoV-19/England/ALDP-B8468F/2020\|EPI_ISL_675204\|20 | ···················· | ···················· | ···················· | ···················· | ···················· | ···················· |
| U | 4866 | hCoV-19/England/ALDP-B83E5B/2020\|EPI_ISL_675205\|20 | ···················· | ···················· | ···················· | ···················· | ···················· | ···················· |
| U | 4867 | hCoV-19/England/ALDP-B8493B/2020\|EPI_ISL_675206\|20 | ···················· | ···················· | ···················· | ···················· | ···················· | ···················· |
| U | 4868 | hCoV-19/England/ALDP-B837E7/2020\|EPI_ISL_675208\|20 | ···················· | ···················· | ···················· | ···················· | ···················· | ···················· |
| U | 4869 | hCoV-19/England/MILK-B8F086/2020\|EPI_ISL_675210\|20 | ···················· | ···················· | ···················· | ···················· | ···················· | ···················· |
| U | 4870 | hCoV-19/England/ALDP-B838C6/2020\|EPI_ISL_675211\|20 | ···················· | ···················· | ···················· | ···················· | ···················· | ···················· |
| U | 4871 | hCoV-19/England/MILK-B8F3AB/2020\|EPI_ISL_675212\|20 | ···················· | ···················· | ···················· | ···················· | ···················· | ···················· |
| U | 4872 | hCoV-19/England/ALDP-B83671/2020\|EPI_ISL_675213\|20 | ···················· | ···················· | ···················· | ···················· | ···················· | ···················· |
| U | 4873 | hCoV-19/England/ALDP-B8387B/2020\|EPI_ISL_675214\|20 | ···················· | ···················· | ···················· | ···················· | ···················· | ···················· |
| U | 4874 | hCoV-19/England/ALDP-B83811/2020\|EPI_ISL_675215\|20 | ···················· | ···················· | ···················· | ···················· | ···················· | ···················· |
| U | 4875 | hCoV-19/England/MILK-B8F04A/2020\|EPI_ISL_675217\|20 | ···················· | ···················· | ···················· | ···················· | ···················· | ···················· |
| U | 4876 | hCoV-19/England/ALDP-B82CAD/2020\|EPI_ISL_675218\|20 | ···················· | ·A·················· | ···················· | ···················· | ···················· | ···A················ |
| U | 4877 | hCoV-19/England/MILK-B8F02C/2020\|EPI_ISL_675219\|20 | ···················· | ···················· | ···················· | ···················· | ···················· | ···················· |
| U | 4878 | hCoV-19/England/ALDP-B837C9/2020\|EPI_ISL_675220\|20 | ···················· | ···················· | ···················· | ···················· | ···················· | ···················· |
| U | 4879 | hCoV-19/England/ALDP-B83AB1/2020\|EPI_ISL_675221\|20 | ···················· | ···················· | ···················· | ···················· | ···················· | ···················· |
| U | 4880 | hCoV-19/England/ALDP-B84959/2020\|EPI_ISL_675222\|20 | ···················· | ···················· | ···················· | ···················· | ······T············· | ···················· |
| U | 4881 | hCoV-19/England/ALDP-B83C06/2020\|EPI_ISL_675223\|20 | ···················· | ···················· | ···················· | ···················· | ···················· | ···················· |
| U | 4882 | hCoV-19/England/MILK-B8F37E/2020\|EPI_ISL_675224\|20 | ···················· | ···················· | ···················· | ···················· | ···················· | ···················· |
| U | 4883 | hCoV-19/England/ALDP-B84009/2020\|EPI_ISL_675225\|20 | ···················· | ···················· | ···················· | ···················· | ···················· | ···················· |
| U | 4884 | hCoV-19/England/ALDP-B82D04/2020\|EPI_ISL_675226\|20 | ···················· | ···················· | ···················· | ···················· | ···················· | ···················· |
| U | 4885 | hCoV-19/England/MILK-B8EFA8/2020\|EPI_ISL_675228\|20 | ···················· | ···················· | ···················· | ···················· | ···················· | ···················· |
| U | 4886 | hCoV-19/England/ALDP-B83FD0/2020\|EPI_ISL_675230\|20 | ···················· | ···················· | ···················· | ···················· | ···················· | ···················· |
| U | 4887 | hCoV-19/England/ALDP-B84986/2020\|EPI_ISL_675231\|20 | ···················· | ···················· | ···················· | ···················· | ···················· | ···················· |
| U | 4888 | hCoV-19/England/ALDP-B85B16/2020\|EPI_ISL_675232\|20 | ···················· | ···················· | ···················· | ···················· | ···················· | ···················· |
| U | 4889 | hCoV-19/England/ALDP-B83ED3/2020\|EPI_ISL_675233\|20 | ···················· | ···················· | ···················· | ···················· | ······T············· | ···················· |
| U | 4890 | hCoV-19/England/ALDP-B842C7/2020\|EPI_ISL_675235\|20 | ···················· | ···················· | ···················· | ···················· | ···················· | ···················· |
| U | 4891 | hCoV-19/England/ALDP-B83C24/2020\|EPI_ISL_675237\|20 | ···················· | ···················· | ···················· | ···················· | ···················· | ···················· |
| U | 4892 | hCoV-19/England/MILK-B8EDBD/2020\|EPI_ISL_675238\|20 | ···················· | ···················· | ···················· | ···················· | ···················· | ···················· |
| U | 4893 | hCoV-19/England/ALDP-B82D9B/2020\|EPI_ISL_675239\|20 | ···················· | ···················· | ···················· | ···················· | ···················· | ···················· |
| U | 4894 | hCoV-19/England/ALDP-B841E8/2020\|EPI_ISL_675240\|20 | ···················· | ···················· | ···················· | ···················· | ···················· | ···················· |
| U | 4895 | hCoV-19/England/MILK-B8F00E/2020\|EPI_ISL_675241\|20 | ···················· | ···················· | ···················· | ···················· | ···················· | ···················· |
| U | 4896 | hCoV-19/England/MILK-B8EE32/2020\|EPI_ISL_675243\|20 | ···················· | ·A·················· | ···················· | ···················· | ···················· | ···A················ |
| U | 4897 | hCoV-19/England/MILK-B8EF20/2020\|EPI_ISL_675244\|20 | ···················· | ···················· | ···················· | ···················· | ···················· | ···················· |
| U | 4898 | hCoV-19/England/ALDP-B83802/2020\|EPI_ISL_675245\|20 | ···················· | ···················· | ···················· | ···················· | ···················· | ···················· |
| U | 4899 | hCoV-19/England/ALDP-B841BB/2020\|EPI_ISL_675246\|20 | ···················· | ···················· | ···················· | ···················· | ···················· | ···················· |
| U | 4900 | hCoV-19/England/ALDP-B845CE/2020\|EPI_ISL_675248\|20 | ···················· | ···················· | ···················· | ···················· | ···················· | ···················· |
| U | 4901 | hCoV-19/England/ALDP-B829A6/2020\|EPI_ISL_675250\|20 | ···················· | ···················· | ···················· | ···················· | ···················· | ········R··········· |
| U | 4902 | hCoV-19/England/ALDP-B83680/2020\|EPI_ISL_675251\|20 | ···················· | ···················· | ···················· | ···················· | ···················· | ···················· |
| U | 4903 | hCoV-19/England/ALDP-B8476E/2020\|EPI_ISL_675252\|20 | ···················· | ·A·················· | ···················· | ···················· | ···················· | ···················· |
| U | 4904 | hCoV-19/England/ALDP-B8574F/2020\|EPI_ISL_675253\|20 | ···················· | ···················· | ···················· | ···················· | ···················· | ···················· |
| U | 4905 | hCoV-19/England/ALDP-B835ED/2020\|EPI_ISL_675254\|20 | ···················· | ···················· | ···················· | ···················· | ···················· | ···A················ |
| U | 4906 | hCoV-19/England/ALDP-B84625/2020\|EPI_ISL_675255\|20 | ···················· | ···················· | ···················· | ···················· | ···················· | ···················· |
| U | 4907 | hCoV-19/England/ALDP-B8576D/2020\|EPI_ISL_675256\|20 | ···················· | ···················· | ···················· | ···················· | ···················· | ···················· |
| U | 4908 | hCoV-19/England/ALDP-B83BEB/2020\|EPI_ISL_675257\|20 | ···················· | ···················· | ···················· | ···················· | ···················· | ···················· |

| | | | | | | | | |
|---|---|---|---|---|---|---|---|---|
| U | 4909 | hCoV-19/England/ALDP-B857C7/2020\|EPI_ISL_675258\|20 | ···················· | ···················· | ····················· | ···················· | ···················· | ···················· |
| U | 4910 | hCoV-19/England/ALDP-B85712/2020\|EPI_ISL_675259\|20 | ···················· | ···················· | ····················· | ···················· | ···················· | ···················· |
| U | 4911 | hCoV-19/England/MILK-B8F2CC/2020\|EPI_ISL_675260\|20 | ···················· | ···················· | ····················· | ···················· | ···················· | ···················· |
| U | 4912 | hCoV-19/England/ALDP-B84564/2020\|EPI_ISL_675261\|20 | ···················· | ···················· | ····················· | ···················· | ···················· | ···················· |
| U | 4913 | hCoV-19/England/ALDP-B8394B/2020\|EPI_ISL_675262\|20 | ···················· | ···················· | ····················· | ···················· | ···················· | ···················· |
| U | 4914 | hCoV-19/England/ALDP-B829E2/2020\|EPI_ISL_675263\|20 | ···················· | ···················· | ····················· | ···················· | ···················· | ···················· |
| U | 4915 | hCoV-19/England/MILK-B8F2DB/2020\|EPI_ISL_675264\|20 | ···················· | ···················· | ····················· | ···················· | ···················· | ···················· |
| U | 4916 | hCoV-19/England/ALDP-B82CF8/2020\|EPI_ISL_675265\|20 | ···················· | ···················· | ····················· | ···················· | ···················· | ···················· |
| U | 4917 | hCoV-19/England/ALDP-B842B8/2020\|EPI_ISL_675266\|20 | ···················· | ···················· | ····················· | ···················· | ···················· | ···················· |
| U | 4918 | hCoV-19/England/ALDP-B8385D/2020\|EPI_ISL_675268\|20 | ···················· | ···················· | ····················· | ···················· | ···················· | ···················· |
| U | 4919 | hCoV-19/England/ALDP-B8479B/2020\|EPI_ISL_675269\|20 | ···················· | ···················· | ····················· | ···················· | ···················· | ···················· |
| U | 4920 | hCoV-19/England/ALDP-B83653/2020\|EPI_ISL_675270\|20 | ···················· | ···················· | ····················· | ···················· | ···················· | ···················· |
| U | 4921 | hCoV-19/England/ALDP-B836BD/2020\|EPI_ISL_675271\|20 | ···················· | ···················· | ····················· | ···················· | ···················· | ···················· |
| U | 4922 | hCoV-19/England/ALDP-B82DC8/2020\|EPI_ISL_675272\|20 | ···················· | ···················· | ····················· | ···················· | ···················· | ···················· |
| U | 4923 | hCoV-19/England/ALDP-B83FFE/2020\|EPI_ISL_675273\|20 | ···················· | ···················· | ····················· | ···················· | ···················· | ···················· |
| U | 4924 | hCoV-19/England/ALDP-B82F86/2020\|EPI_ISL_675274\|20 | ···················· | ·········T·········· | ····················· | ···················· | ···················· | ···················· |
| U | 4925 | hCoV-19/England/MILK-B8F323/2020\|EPI_ISL_675275\|20 | ···················· | ···················· | ····················· | ···················· | ···················· | ···················· |
| U | 4926 | hCoV-19/England/MILK-B8F226/2020\|EPI_ISL_675276\|20 | ···················· | ···················· | ····················· | ···················· | ···················· | ···················· |
| U | 4927 | hCoV-19/England/MILK-B8F059/2020\|EPI_ISL_675277\|20 | ···················· | ···················· | ····················· | ···················· | ···················· | ···················· |
| U | 4928 | hCoV-19/England/ALDP-B83EA6/2020\|EPI_ISL_675278\|20 | ···················· | ···················· | ····················· | ···················· | ···················· | ···················· |
| U | 4929 | hCoV-19/England/ALDP-B837BA/2020\|EPI_ISL_675279\|20 | ···················· | ···················· | ····················· | ···················· | ···················· | ···················· |
| U | 4930 | hCoV-19/England/ALDP-B83A57/2020\|EPI_ISL_675280\|20 | ···················· | ···················· | ····················· | ···················· | ···················· | ···················· |
| U | 4931 | hCoV-19/England/ALDP-B844B2/2020\|EPI_ISL_675281\|20 | ···················· | ···················· | ····················· | ···················· | ···················· | ···················· |
| U | 4932 | hCoV-19/England/MILK-B8F402/2020\|EPI_ISL_675282\|20 | ···················· | ···················· | ····················· | ···················· | ···················· | ···················· |
| U | 4933 | hCoV-19/England/ALDP-B8425E/2020\|EPI_ISL_675283\|20 | ···················· | ···················· | ····················· | ···················· | ···················· | ···················· |
| U | 4934 | hCoV-19/England/MILK-B8F147/2020\|EPI_ISL_675284\|20 | ···················· | ·A·················· | ····················· | ···················· | ···················· | ···A················ |
| U | 4935 | hCoV-19/England/MILK-B8EE41/2020\|EPI_ISL_675285\|20 | ···················· | ···················· | ····················· | ···················· | ···················· | ···················· |
| U | 4936 | hCoV-19/England/ALDP-B84151/2020\|EPI_ISL_675287\|20 | ···················· | ···················· | ····················· | ···················· | ···················· | ···················· |
| U | 4937 | hCoV-19/England/ALDP-B83741/2020\|EPI_ISL_675288\|20 | ···················· | ···················· | ····················· | ···················· | ···················· | ···················· |
| U | 4938 | hCoV-19/England/ALDP-B838A8/2020\|EPI_ISL_675300\|20 | ···················· | ···················· | ····················· | ···················· | ···················· | ···················· |
| U | 4939 | hCoV-19/England/MILK-B8F217/2020\|EPI_ISL_675301\|20 | ···················· | ···················· | ····················· | ···················· | ···················· | ···················· |
| U | 4940 | hCoV-19/England/ALDP-B8431F/2020\|EPI_ISL_675302\|20 | ···················· | ···················· | ····················· | ···················· | ···················· | ···················· |
| U | 4941 | hCoV-19/England/ALDP-B857F4/2020\|EPI_ISL_675303\|20 | ···················· | ···················· | ····················· | ···················· | ···················· | ···················· |
| U | 4942 | hCoV-19/England/MILK-B8F2F9/2020\|EPI_ISL_675305\|20 | ···················· | ···················· | ····················· | ···················· | ···················· | ···················· |
| U | 4943 | hCoV-19/England/MILK-B8EDF9/2020\|EPI_ISL_675306\|20 | ···················· | ···················· | ····················· | ···················· | ···················· | ···················· |
| U | 4944 | hCoV-19/England/MILK-B8EF3F/2020\|EPI_ISL_675307\|20 | ···················· | ···················· | ····················· | ···················· | ···················· | ···················· |
| U | 4945 | hCoV-19/England/MILK-B8F305/2020\|EPI_ISL_675308\|20 | ···················· | ···················· | ····················· | ···················· | ···················· | ···················· |
| U | 4946 | hCoV-19/England/ALDP-B82A3A/2020\|EPI_ISL_675310\|20 | ···················· | ···················· | ····················· | ···················· | ···················· | ···A················ |
| U | 4947 | hCoV-19/England/MILK-B8EF6C/2020\|EPI_ISL_675311\|20 | ···················· | ···················· | ····················· | ···················· | ···················· | ···················· |
| U | 4948 | hCoV-19/England/ALDP-B83644/2020\|EPI_ISL_675312\|20 | ···················· | ···················· | ····················· | ···················· | ···················· | ···················· |
| U | 4949 | hCoV-19/England/MILK-B8F077/2020\|EPI_ISL_675313\|20 | ···················· | ···················· | ····················· | ···················· | ···················· | ···················· |
| U | 4950 | hCoV-19/Sweden/20-53303/2020\|EPI_ISL_676535\|2020-1 | ···················· | ···················· | ····················· | ···················· | ······T············· | ···················· |
| U | 4951 | hCoV-19/Sweden/20-53325/2020\|EPI_ISL_676536\|2020-1 | ···················· | ·A·················· | ····················· | ···················· | ···················· | ···A················ |
| U | 4952 | hCoV-19/Sweden/20-53328/2020\|EPI_ISL_676537\|2020-1 | ···················· | ···················· | ····················· | ···················· | ···················· | ···················· |
| U | 4953 | hCoV-19/Sweden/20-53329/2020\|EPI_ISL_676538\|2020-1 | ···················· | ···················· | ····················· | ···················· | ···················· | ···················· |
| U | 4954 | hCoV-19/Sweden/20-53378/2020\|EPI_ISL_676539\|2020-1 | ···················· | ···················· | ····················· | ···················· | ···················· | ···················· |
| U | 4955 | hCoV-19/Sweden/20-53382/2020\|EPI_ISL_676540\|2020-1 | ···················· | ···················· | ····················· | ···················· | ···················· | ···················· |
| U | 4956 | hCoV-19/France/ARA-22140/2020\|EPI_ISL_676542\|2020- | ···················· | ···················· | ····················· | ···················· | ···················· | ···················· |
| U | 4957 | hCoV-19/France/ARA-24293/2020\|EPI_ISL_676543\|2020- | ···················· | ···················· | ····················· | ···················· | ···················· | ···················· |
| U | 4958 | hCoV-19/France/ARA-24305/2020\|EPI_ISL_676544\|2020- | ···················· | ···················· | ····················· | ···················· | ···················· | ···················· |
| U | 4959 | hCoV-19/France/ARA-24352/2020\|EPI_ISL_676545\|2020- | ···················· | ···················· | ····················· | ···················· | ···················· | ···················· |
| U | 4960 | hCoV-19/France/ARA-24445/2020\|EPI_ISL_676546\|2020- | ···················· | ···················· | ····················· | ···················· | ···················· | ···················· |
| U | 4961 | hCoV-19/France/ARA-24473/2020\|EPI_ISL_676547\|2020- | ···················· | ···················· | ····················· | ···················· | ···················· | ···················· |
| U | 4962 | hCoV-19/France/ARA-24538/2020\|EPI_ISL_676548\|2020- | ···················· | ···················· | ····················· | ···················· | ···················· | ···················· |
| U | 4963 | hCoV-19/France/ARA-24895/2020\|EPI_ISL_676550\|2020- | ···················· | ···················· | ····················· | ···················· | ···················· | ···················· |
| U | 4964 | hCoV-19/France/ARA-24935/2020\|EPI_ISL_676551\|2020- | ···················· | ···················· | ····················· | ···················· | ···················· | ···················· |
| U | 4965 | hCoV-19/France/ARA-25111/2020\|EPI_ISL_676552\|2020- | ···················· | ···················· | ····················· | ···················· | ···················· | ···················· |
| U | 4966 | hCoV-19/France/ARA-25131/2020\|EPI_ISL_676553\|2020- | ···················· | ···················· | ····················· | ···················· | ···················· | ···················· |
| U | 4967 | hCoV-19/France/ARA-25219/2020\|EPI_ISL_676554\|2020- | ···················· | ···················· | ····················· | ···················· | ···················· | ···················· |
| U | 4968 | hCoV-19/France/ARA-25382/2020\|EPI_ISL_676555\|2020- | ···················· | ···················· | ····················· | ···················· | ···················· | ···················· |
| U | 4969 | hCoV-19/France/ARA-25403/2020\|EPI_ISL_676556\|2020- | ···················· | ···················· | ····················· | ···················· | ···················· | ···················· |
| U | 4970 | hCoV-19/France/ARA-25424/2020\|EPI_ISL_676557\|2020- | ···················· | ···················· | ····················· | ···················· | ···················· | ···················· |
| U | 4971 | hCoV-19/France/ARA-26054/2020\|EPI_ISL_676558\|2020- | ···················· | ···················· | ····················· | ···················· | ···················· | ···················· |
| U | 4972 | hCoV-19/France/ARA-26087/2020\|EPI_ISL_676559\|2020- | ···················· | ···················· | ····················· | ···················· | ···················· | ···················· |
| U | 4973 | hCoV-19/France/ARA-26157/2020\|EPI_ISL_676560\|2020- | ···················· | ···················· | ····················· | ···················· | ···················· | ···················· |
| U | 4974 | hCoV-19/France/ARA-26174/2020\|EPI_ISL_676561\|2020- | ···················· | ···················· | ····················· | ···················· | ···················· | ···················· |
| U | 4975 | hCoV-19/France/ARA-26417/2020\|EPI_ISL_676562\|2020- | ···················· | ···················· | ····················· | ···················· | ···················· | ···················· |
| U | 4976 | hCoV-19/France/ARA-26541/2020\|EPI_ISL_676563\|2020- | ···················· | ···················· | ····················· | ···················· | ···················· | ···················· |
| U | 4977 | hCoV-19/France/ARA-26693/2020\|EPI_ISL_676564\|2020- | ···················· | ···················· | ····················· | ···················· | ···················· | ···················· |
| U | 4978 | hCoV-19/France/ARA-26824/2020\|EPI_ISL_676565\|2020- | ···················· | ···················· | ····················· | ···················· | ···················· | ···················· |
| U | 4979 | hCoV-19/France/ARA-27101/2020\|EPI_ISL_676566\|2020- | ···················· | ···················· | ····················· | ···················· | ···················· | ···················· |
| U | 4980 | hCoV-19/France/ARA-27109/2020\|EPI_ISL_676567\|2020- | ···················· | ···················· | ····················· | ···················· | ···················· | ···················· |
| U | 4981 | hCoV-19/France/ARA-27161/2020\|EPI_ISL_676568\|2020- | ···················· | ···················· | ···C················· | ···················· | ···················· | ···················· |
| U | 4982 | hCoV-19/France/ARA-27275/2020\|EPI_ISL_676569\|2020- | ···················· | ···················· | ····················· | ···················· | ···················· | ···················· |
| U | 4983 | hCoV-19/France/ARA-27286/2020\|EPI_ISL_676570\|2020- | ···················· | ···················· | ····················· | ···················· | ···················· | ···················· |
| U | 4984 | hCoV-19/France/ARA-27306/2020\|EPI_ISL_676571\|2020- | ···················· | ···················· | ····················· | ···················· | ···················· | ···················· |
| U | 4985 | hCoV-19/France/ARA-27314/2020\|EPI_ISL_676572\|2020- | ···················· | ···················· | ····················· | ···················· | ···················· | ···················· |
| U | 4986 | hCoV-19/France/ARA-27377/2020\|EPI_ISL_676573\|2020- | ···················· | ···················· | ····················· | ···················· | ···················· | ···················· |
| U | 4987 | hCoV-19/Serbia/Subotica-016620604/2020\|EPI_ISL_676 | ···················· | ···················· | ····················· | ···················· | ···················· | ···················· |
| U | 4988 | hCoV-19/Serbia/Sremska Mitrovica-011821207/2020\|EP | ···················· | ···················· | ····················· | ···················· | ···················· | ···················· |

| | | | | | | | | |
|---|---|---|---|---|---|---|---|---|
| U | 4989 | hCoV-19/Serbia/Pancevo-007910104/2020\|EPI_ISL_6765 | ···················· | ···················· | ···················· | ···················· | ···················· | ···················· |
| U | 4990 | hCoV-19/Serbia/Backa Topola-00V720304/2020\|EPI_ISL | ···················· | ···················· | ···················· | ···················· | ···················· | ···················· |
| U | 4991 | hCoV-19/Serbia/Novi Sad-091180205/2020\|EPI_ISL_676 | ···················· | ···················· | ···················· | ···················· | ···················· | ···················· |
| U | 4992 | hCoV-19/Serbia/Sremska Mitrovica-010500907/2020\|EP | ···················· | ···················· | ···················· | ···················· | ···················· | ···················· |
| U | 4993 | hCoV-19/Serbia/Novi Sad-004460704/2020\|EPI_ISL_676 | ···················· | ···················· | ···················· | ···················· | ···················· | ···················· |
| U | 4994 | hCoV-19/Serbia/Sremska Mitrovica-022620904/2020\|EP | ···················· | ···················· | ···················· | ···················· | ···················· | ···················· |
| U | 4995 | hCoV-19/Serbia/Novi Sad-210722506/2020\|EPI_ISL_676 | ···················· | ···················· | ···················· | ···················· | ···················· | ···················· |
| U | 4996 | hCoV-19/Serbia/Novi Sad-212182506/2020\|EPI_ISL_676 | ···················· | ···················· | ···················· | ···················· | ···················· | ···················· |
| U | 4997 | hCoV-19/Serbia/Novi Sad-212282506/2020\|EPI_ISL_676 | ···················· | ···················· | ···················· | ···················· | ···················· | ···················· |
| U | 4998 | hCoV-19/Serbia/Subotica-101360605/2020\|EPI_ISL_676 | ···················· | ···················· | ···················· | ···················· | ···················· | ···················· |
| U | 4999 | hCoV-19/Serbia/Pancevo-101740605/2020\|EPI_ISL_6765 | ···················· | ···················· | ···················· | ···················· | ···················· | ···················· |
| U | 5000 | hCoV-19/Serbia/Vrsac-101760605/2020\|EPI_ISL_676587 | ···················· | ···················· | ···················· | ···················· | ···················· | ···················· |
| U | 5001 | hCoV-19/Serbia/Novi Sad-111681105/2020\|EPI_ISL_676 | ···················· | ···················· | ···················· | ···················· | ···················· | ···················· |
| U | 5002 | hCoV-19/Serbia/Subotica-125991705/2020\|EPI_ISL_676 | ···················· | ···················· | ···················· | ···················· | ···················· | ···················· |
| U | 5003 | hCoV-19/Serbia/Novi Sad-158472905/2020\|EPI_ISL_676 | ···················· | ···················· | ···················· | ···················· | ···················· | ···················· |
| U | 5004 | hCoV-19/Serbia/Novi Sad-158502905/2020\|EPI_ISL_676 | ···················· | ···················· | ···················· | ···················· | ···················· | ···················· |
| U | 5005 | hCoV-19/Serbia/Novi Sad-196221606/2020\|EPI_ISL_676 | ···················· | ···················· | ···················· | ···················· | ···················· | ···················· |
| U | 5006 | hCoV-19/Serbia/Zrenjanin-033961404/2020\|EPI_ISL_67 | ···················· | ···················· | ···················· | ···················· | ···················· | ···················· |
| U | 5007 | hCoV-19/Serbia/Pancevo-037181504/2020\|EPI_ISL_6765 | ···················· | ···················· | ···················· | ···················· | ···················· | ···················· |
| U | 5008 | hCoV-19/Serbia/Vrsac-040471604/2020\|EPI_ISL_676595 | ···················· | ···················· | ···················· | ···················· | ···················· | ···················· |
| U | 5009 | hCoV-19/Serbia/Srbobran-043051704/2020\|EPI_ISL_676 | ···················· | ···················· | ···················· | ···················· | ···················· | ···················· |
| U | 5010 | hCoV-19/Serbia/Titel-054312104/2020\|EPI_ISL_676597 | ···················· | ···················· | ···················· | ···················· | ···················· | ···················· |
| U | 5011 | hCoV-19/Serbia/Titel-059642304/2020\|EPI_ISL_676598 | ···················· | ···················· | ···················· | ···················· | ···················· | ···················· |
| U | 5012 | hCoV-19/Serbia/Subotica-062172304/2020\|EPI_ISL_676 | ···················· | ···················· | ···················· | ···················· | ···················· | ···················· |
| U | 5013 | hCoV-19/Serbia/Pancevo-075362804/2020\|EPI_ISL_6766 | ···················· | ···················· | ···················· | ···················· | ···················· | ···················· |
| U | 5014 | hCoV-19/Serbia/Novi Sad-196981606/2020\|EPI_ISL_676 | ···················· | ···················· | ···················· | ···················· | ···················· | ···················· |
| U | 5015 | hCoV-19/Serbia/Novi Sad-022850904/2020\|EPI_ISL_676 | ···················· | ···················· | ···················· | ···················· | ···················· | ···················· |
| U | 5016 | hCoV-19/Serbia/Novi Sad-023010904/2020\|EPI_ISL_676 | ···················· | ···················· | ···················· | ···················· | ···················· | ···················· |
| U | 5017 | hCoV-19/USA/TX-DSHS-1451/2020\|EPI_ISL_676642\|2020- | ···················· | ···················· | ···················· | ···················· | ···················· | ···················· |
| U | 5018 | hCoV-19/USA/TX-DSHS-1462/2020\|EPI_ISL_676643\|2020- | ···················· | ···················· | ···················· | ···················· | ···················· | ···················· |
| U | 5019 | hCoV-19/USA/TX-DSHS-1468/2020\|EPI_ISL_676644\|2020- | ···················· | ·A·················· | ···················· | ···················· | ···················· | ···················· |
| U | 5020 | hCoV-19/USA/CA-IGI-0673/2020\|EPI_ISL_677866\|2020-0 | ···················· | ···················· | ···················· | ···················· | ···················· | ···················· |
| U | 5021 | hCoV-19/USA/CA-IGI-0676/2020\|EPI_ISL_677867\|2020-0 | ···················· | ···················· | ···················· | ···················· | ···················· | ···················· |
| U | 5022 | hCoV-19/USA/CA-IGI-0684/2020\|EPI_ISL_677868\|2020-0 | ···················· | ·A·················· | ···················· | ···················· | ···················· | ···················· |
| U | 5023 | hCoV-19/USA/CA-IGI-0686/2020\|EPI_ISL_677869\|2020-0 | ···················· | ·A·················· | ···················· | ···················· | ···················· | ···················· |
| U | 5024 | hCoV-19/USA/CA-IGI-0687/2020\|EPI_ISL_677870\|2020-0 | ···················· | ···················· | ···················· | ···················· | ···················· | ···················· |
| U | 5025 | hCoV-19/USA/CA-IGI-0688/2020\|EPI_ISL_677871\|2020-0 | ···················· | ···················· | ···················· | ···················· | ········A··········· | ···················· |
| U | 5026 | hCoV-19/USA/CA-IGI-0691/2020\|EPI_ISL_677872\|2020-0 | ···················· | ···················· | ···················· | ···················· | ···················· | ···················· |
| U | 5027 | hCoV-19/USA/CA-IGI-0692/2020\|EPI_ISL_677873\|2020-0 | ···················· | ···················· | ···················· | ···················· | ···················· | ···················· |
| U | 5028 | hCoV-19/USA/CA-IGI-0693/2020\|EPI_ISL_677874\|2020-0 | ···················· | ···················· | ···················· | ···················· | ···················· | ···················· |
| U | 5029 | hCoV-19/USA/CA-IGI-0700/2020\|EPI_ISL_677875\|2020-0 | ···················· | ···················· | ···················· | ···················· | ···················· | ···················· |
| U | 5030 | hCoV-19/USA/CA-IGI-0703/2020\|EPI_ISL_677876\|2020-0 | ···················· | ···················· | ···················· | ···················· | ···················· | ···················· |
| U | 5031 | hCoV-19/USA/CA-IGI-0710/2020\|EPI_ISL_677877\|2020-0 | ···················· | ···················· | ···················· | ···················· | ···················· | ···················· |
| U | 5032 | hCoV-19/USA/CA-IGI-0723/2020\|EPI_ISL_677878\|2020-0 | ···················· | ···················· | ···················· | ···················· | ···················· | ···················· |
| U | 5033 | hCoV-19/USA/CA-IGI-0725/2020\|EPI_ISL_677879\|2020-0 | ···················· | ···················· | ···················· | ···················· | ···················· | ···················· |
| U | 5034 | hCoV-19/USA/CA-IGI-0730/2020\|EPI_ISL_677880\|2020-0 | ···················· | ···················· | ···················· | ···················· | ···················· | ···················· |
| U | 5035 | hCoV-19/USA/CA-IGI-0740/2020\|EPI_ISL_677883\|2020-0 | ···················· | ···················· | ···················· | ···················· | ········A··········· | ···················· |
| U | 5036 | hCoV-19/USA/CA-IGI-0741/2020\|EPI_ISL_677884\|2020-0 | ···················· | ···················· | ···················· | ···················· | ···················· | ···················· |
| U | 5037 | hCoV-19/USA/CA-IGI-0760/2020\|EPI_ISL_677885\|2020-0 | ···················· | ···················· | ···················· | ···················· | ···················· | ···················· |
| U | 5038 | hCoV-19/USA/CA-IGI-0765/2020\|EPI_ISL_677886\|2020-0 | ···················· | ···················· | ···················· | ···················· | ···················· | ···················· |
| U | 5039 | hCoV-19/USA/CA-IGI-0769/2020\|EPI_ISL_677887\|2020-0 | ···················· | ···················· | ···················· | ···················· | ···················· | ···················· |
| U | 5040 | hCoV-19/USA/CA-IGI-0793/2020\|EPI_ISL_677889\|2020-0 | ···················· | ···················· | ···················· | ···················· | ···················· | ···················· |
| U | 5041 | hCoV-19/USA/CA-IGI-0796/2020\|EPI_ISL_677890\|2020-0 | ···················· | ···················· | ···················· | ···················· | ···················· | ···················· |
| U | 5042 | hCoV-19/USA/CA-IGI-0797/2020\|EPI_ISL_677891\|2020-0 | ···················· | ···················· | ···················· | ···················· | ···················· | ···················· |
| U | 5043 | hCoV-19/USA/CA-IGI-0799/2020\|EPI_ISL_677892\|2020-0 | ···················· | ···················· | ···················· | ···················· | ···················· | ···················· |
| U | 5044 | hCoV-19/USA/CA-IGI-0800/2020\|EPI_ISL_677893\|2020-0 | ···················· | ···················· | ···················· | ···················· | ···················· | ···················· |
| U | 5045 | hCoV-19/USA/CA-IGI-0801/2020\|EPI_ISL_677894\|2020-0 | ···················· | ···················· | ···················· | ···················· | ···················· | ···················· |
| U | 5046 | hCoV-19/USA/CA-IGI-0808/2020\|EPI_ISL_677896\|2020-0 | ···················· | ···················· | ···················· | ···················· | ···················· | ···················· |
| U | 5047 | hCoV-19/USA/CA-IGI-0810/2020\|EPI_ISL_677897\|2020-0 | ···················· | ···················· | ···················· | ···················· | ···················· | ···················· |
| U | 5048 | hCoV-19/USA/CA-IGI-0813/2020\|EPI_ISL_677898\|2020-0 | ···················· | ···················· | ···················· | ···················· | ···················· | ···················· |
| U | 5049 | hCoV-19/USA/CA-IGI-0815/2020\|EPI_ISL_677899\|2020-0 | ···················· | ···················· | ···················· | ···················· | ···················· | ···················· |
| U | 5050 | hCoV-19/USA/CA-IGI-0816/2020\|EPI_ISL_677900\|2020-0 | ···················· | ···················· | ···················· | ···················· | ···················· | ···················· |
| U | 5051 | hCoV-19/USA/CA-IGI-0817/2020\|EPI_ISL_677901\|2020-0 | ···················· | ···················· | ···················· | ···················· | ···················· | ···················· |
| U | 5052 | hCoV-19/USA/CA-IGI-0826/2020\|EPI_ISL_677904\|2020-0 | ···················· | ···················· | ···················· | ···················· | ···················· | ···················· |
| U | 5053 | hCoV-19/USA/CA-IGI-0830/2020\|EPI_ISL_677905\|2020-0 | ···················· | ···················· | ···················· | ···················· | ···················· | ···················· |
| U | 5054 | hCoV-19/USA/CA-IGI-0836/2020\|EPI_ISL_677907\|2020-0 | ···················· | ···················· | ···················· | ···················· | ···················· | ···················· |
| U | 5055 | hCoV-19/Saudi Arabia/KAUST-JEDDAH1291/2020\|EPI_ISL | ···················· | ···················· | ···················· | ···················· | ···················· | ···················· |
| U | 5056 | hCoV-19/Saudi Arabia/KAUST-JEDDAH1292/2020\|EPI_ISL | ···················· | ···················· | ···················· | ···················· | ···················· | ···················· |
| U | 5057 | hCoV-19/Saudi Arabia/KAUST-JEDDAH1295/2020\|EPI_ISL | ···················· | ·A·················· | ···················· | ···················· | ···················· | ···················· |
| U | 5058 | hCoV-19/Saudi Arabia/KAUST-JEDDAH1296/2020\|EPI_ISL | ···················· | ···················· | ···················· | ···················· | ···················· | ···················· |
| U | 5059 | hCoV-19/Saudi Arabia/KAUST-JEDDAH1299/2020\|EPI_ISL | ···················· | ···················· | ···················· | ···················· | ···················· | ···················· |
| U | 5060 | hCoV-19/Saudi Arabia/KAUST-JEDDAH1301/2020\|EPI_ISL | ···················· | ···················· | ···················· | ···················· | ···················· | ···················· |
| U | 5061 | hCoV-19/Saudi Arabia/KAUST-JEDDAH1316/2020\|EPI_ISL | ···················· | ···················· | ···················· | ···················· | ···················· | ···················· |
| U | 5062 | hCoV-19/Saudi Arabia/KAUST-JEDDAH1317/2020\|EPI_ISL | ···················· | ···················· | ···················· | ···················· | ···················· | ···················· |
| U | 5063 | hCoV-19/Saudi Arabia/KAUST-JEDDAH1318/2020\|EPI_ISL | ···················· | ···················· | ···················· | ···················· | ···················· | ···················· |
| U | 5064 | hCoV-19/Saudi Arabia/KAUST-JEDDAH1319/2020\|EPI_ISL | ···················· | ···················· | ···················· | ···················· | ···················· | ···················· |
| U | 5065 | hCoV-19/Saudi Arabia/KAUST-JEDDAH1325/2020\|EPI_ISL | ···················· | ···················· | ···················· | ···················· | ···················· | ···················· |
| U | 5066 | hCoV-19/Saudi Arabia/KAUST-JEDDAH1333/2020\|EPI_ISL | ···················· | ·A·················· | ···················· | ···················· | ···················· | ··A················· |
| U | 5067 | hCoV-19/Saudi Arabia/KAUST-JEDDAH1337/2020\|EPI_ISL | ···················· | ···················· | ···················· | ···················· | ···················· | ···················· |
| U | 5068 | hCoV-19/Saudi Arabia/KAUST-JEDDAH1338/2020\|EPI_ISL | ···················· | ···················· | ···················· | ···················· | ···················· | ···················· |

| | | | | | | | |
|---|---|---|---|---|---|---|---|
| U | 5069 hCoV-19/Saudi Arabia/KAUST-JEDDAH1339/2020\|EPI_ISL | ···················· | ·A·················· | ···················· | ···················· | ···················· | ···A················ |
| U | 5070 hCoV-19/Saudi Arabia/KAUST-JEDDAH500/2020\|EPI_ISL_ | ···················· | ···················· | ···················· | ···················· | ···················· | ···················· |
| U | 5071 hCoV-19/Saudi Arabia/KAUST-JEDDAH533/2020\|EPI_ISL_ | ···················· | ···················· | ···················· | ···················· | ···················· | ···················· |
| U | 5072 hCoV-19/Saudi Arabia/KAUST-JEDDAH560/2020\|EPI_ISL_ | ···················· | ···················· | ···················· | ···················· | ···················· | ···················· |
| U | 5073 hCoV-19/Saudi Arabia/KAUST-JEDDAH63/2020\|EPI_ISL_6 | ···················· | ·A·················· | ···················· | ···················· | ···················· | ···················· |
| U | 5074 hCoV-19/Saudi Arabia/KAUST-JEDDAH680/2020\|EPI_ISL_ | ···················· | ···················· | ···················· | ···················· | ···················· | ···················· |
| U | 5075 hCoV-19/Saudi Arabia/KAUST-JEDDAH682/2020\|EPI_ISL_ | ···················· | ·A·················· | ···················· | ···················· | ···················· | ···A················ |
| U | 5076 hCoV-19/Saudi Arabia/KAUST-JEDDAH719/2020\|EPI_ISL_ | ···················· | ···················· | ···················· | ···················· | ···················· | ···················· |
| U | 5077 hCoV-19/Saudi Arabia/KAUST-JEDDAH742/2020\|EPI_ISL_ | ···················· | ···················· | ···················· | ···················· | ···················· | ···················· |
| U | 5078 hCoV-19/Saudi Arabia/KAUST-MADINAH259/2020\|EPI_ISL | ···················· | ···················· | ···················· | ···················· | ···················· | ···················· |
| U | 5079 hCoV-19/Saudi Arabia/KAUST-MADINAH1482/2020\|EPI_IS | ···················· | ·A·················· | ···················· | ···················· | ···················· | ···A················ |
| U | 5080 hCoV-19/Saudi Arabia/KAUST-MADINAH1101/2020\|EPI_IS | ···················· | ···················· | ···················· | ···················· | ···················· | ···················· |
| U | 5081 hCoV-19/Saudi Arabia/KAUST-MADINAH669/2020\|EPI_ISL | ···················· | ···················· | ···················· | ···················· | ···················· | ···················· |
| U | 5082 hCoV-19/Saudi Arabia/KAUST-MADINAH1489/2020\|EPI_IS | ···················· | ·A·················· | ···················· | ···················· | ···················· | ···A················ |
| U | 5083 hCoV-19/Saudi Arabia/KAUST-MADINAH1128/2020\|EPI_IS | ···················· | ···················· | ···················· | ···················· | ···················· | ···················· |
| U | 5084 hCoV-19/Wales/PHWC-48F982/2020\|EPI_ISL_681166\|2020 | ···················· | ···················· | ···················· | ···················· | ···················· | ···················· |
| U | 5087 hCoV-19/Wales/PHWC-48F9CE/2020\|EPI_ISL_681169\|2020 | ···················· | ···················· | ···················· | ···················· | ···················· | ···················· |
| U | 5089 hCoV-19/Wales/PHWC-48F9EC/2020\|EPI_ISL_681171\|2020 | ···················· | ···················· | ···················· | ···················· | ···················· | ···················· |
| U | 5092 hCoV-19/Wales/PHWC-48FA25/2020\|EPI_ISL_681175\|2020 | ···················· | ···················· | ···················· | ···················· | ···················· | ···················· |
| U | 5093 hCoV-19/Wales/PHWC-48FA43/2020\|EPI_ISL_681177\|2020 | ···················· | ···················· | ···················· | ···················· | ···················· | ···················· |
| U | 5094 hCoV-19/Wales/PHWC-48FA70/2020\|EPI_ISL_681180\|2020 | ···················· | ···················· | ···················· | ···················· | ···················· | ···················· |
| U | 5095 hCoV-19/Wales/PHWC-48FA8F/2020\|EPI_ISL_681181\|2020 | ···················· | ···················· | ···················· | ···················· | ···················· | ···················· |
| U | 5096 hCoV-19/Wales/PHWC-48FA9E/2020\|EPI_ISL_681182\|2020 | ···················· | ···················· | ···················· | ···················· | ···················· | ···················· |
| U | 5100 hCoV-19/Wales/PHWC-48FB40/2020\|EPI_ISL_681191\|2020 | ···················· | ···················· | ···················· | ···················· | ···················· | ···················· |
| U | 5101 hCoV-19/Wales/PHWC-48FB5F/2020\|EPI_ISL_681192\|2020 | ···················· | ···················· | ···················· | ···················· | ···················· | ···················· |
| U | 5103 hCoV-19/Wales/PHWC-48FB7D/2020\|EPI_ISL_681194\|2020 | ···················· | ···················· | ···················· | ···················· | ···················· | ···················· |
| U | 5104 hCoV-19/Wales/PHWC-48FB8C/2020\|EPI_ISL_681195\|2020 | ···················· | ···················· | ···················· | ···················· | ···················· | ···················· |
| U | 5105 hCoV-19/Wales/PHWC-48FB9B/2020\|EPI_ISL_681196\|2020 | ···················· | ···················· | ···················· | ···················· | ·····T·············· | ···················· |
| U | 5107 hCoV-19/Wales/PHWC-48FBC8/2020\|EPI_ISL_681199\|2020 | ···················· | ···················· | ···················· | ···················· | ···················· | ···················· |
| U | 5108 hCoV-19/Denmark/DCGC-12223/2020\|EPI_ISL_682404\|202 | ···················· | ···················· | ···················· | ···················· | ···················· | ···················· |
| U | 5109 hCoV-19/Denmark/DCGC-12225/2020\|EPI_ISL_682406\|202 | ···················· | ···················· | ···················· | ···················· | ···················· | ···················· |
| U | 5110 hCoV-19/Denmark/DCGC-12228/2020\|EPI_ISL_682409\|202 | ···················· | ···················· | ···················· | ···················· | ···················· | ···················· |
| U | 5111 hCoV-19/USA/MN-MDH-2101/2020\|EPI_ISL_683730\|2020-1 | ···················· | ···················· | ···················· | ···················· | ···················· | ···················· |
| U | 5112 hCoV-19/USA/MN-MDH-2102/2020\|EPI_ISL_683731\|2020-1 | ···················· | ···················· | ···················· | ···················· | ··NN················ | ···················· |
| U | 5113 hCoV-19/USA/MN-MDH-2103/2020\|EPI_ISL_683732\|2020-1 | ···················· | ···················· | ···················· | ···················· | ···················· | ···················· |
| U | 5114 hCoV-19/USA/MN-MDH-2104/2020\|EPI_ISL_683733\|2020-1 | ···················· | ···················· | ···················· | ···················· | ···················· | ···················· |
| U | 5115 hCoV-19/USA/MN-MDH-2105/2020\|EPI_ISL_683734\|2020-1 | ···················· | ···················· | ···················· | ···················· | ···················· | ···················· |
| U | 5116 hCoV-19/USA/MN-MDH-2106/2020\|EPI_ISL_683735\|2020-1 | ···················· | ···················· | ···················· | ···················· | ···················· | ···················· |
| U | 5117 hCoV-19/USA/MN-MDH-2107/2020\|EPI_ISL_683736\|2020-1 | ···················· | ···················· | ···················· | ···················· | ···················· | ···················· |
| U | 5118 hCoV-19/USA/MN-MDH-2108/2020\|EPI_ISL_683737\|2020-1 | ···················· | ···················· | ···················· | ···················· | ···················· | ···················· |
| U | 5119 hCoV-19/USA/MN-MDH-2109/2020\|EPI_ISL_683738\|2020-1 | ···················· | ···················· | ···················· | ···················· | ···················· | ···················· |
| U | 5120 hCoV-19/USA/MN-MDH-2110/2020\|EPI_ISL_683739\|2020-1 | ···················· | ···················· | ···················· | ···················· | ···················· | ···················· |
| U | 5121 hCoV-19/England/CAMC-B0989B/2020\|EPI_ISL_672971\|20 | ···················· | ···················· | ···················· | ···················· | ···················· | ···················· |
| U | 5122 hCoV-19/England/CAMC-B096CE/2020\|EPI_ISL_672973\|20 | ···················· | ···················· | ···················· | ···················· | ···················· | ···················· |
| U | 5123 hCoV-19/England/CAMC-B09400/2020\|EPI_ISL_672974\|20 | ···················· | ···················· | ···················· | ···················· | ···················· | ···················· |
| U | 5124 hCoV-19/England/CAMC-B096A0/2020\|EPI_ISL_672975\|20 | ···················· | ···················· | ···················· | ···················· | ···················· | ···················· |
| U | 5125 hCoV-19/England/CAMC-B09AB3/2020\|EPI_ISL_672976\|20 | ···················· | ···················· | ···················· | ···················· | ···················· | ···················· |
| U | 5126 hCoV-19/England/CAMC-B0A5A2/2020\|EPI_ISL_672978\|20 | ···················· | ···················· | ···················· | ···················· | ···················· | ···················· |
| U | 5127 hCoV-19/England/CAMC-B093C7/2020\|EPI_ISL_672980\|20 | ···················· | ···················· | ···················· | ···················· | ···················· | ···················· |
| U | 5128 hCoV-19/England/CAMC-B09DAB/2020\|EPI_ISL_672981\|20 | ···················· | ···················· | ···················· | ···················· | ···················· | ···················· |
| U | 5129 hCoV-19/England/CAMC-B097AD/2020\|EPI_ISL_672982\|20 | ···················· | ···················· | ···················· | ···················· | ···················· | ···················· |
| U | 5130 hCoV-19/England/CAMC-B08DCA/2020\|EPI_ISL_672983\|20 | ···················· | ···················· | ···················· | ···················· | ···················· | ···················· |
| U | 5131 hCoV-19/England/CAMC-B0A4D2/2020\|EPI_ISL_672984\|20 | ···················· | ···················· | ···················· | ···················· | ···················· | ···················· |
| U | 5132 hCoV-19/England/CAMC-B09190/2020\|EPI_ISL_672985\|20 | ···················· | ···················· | ···················· | ···················· | ···················· | ···················· |
| U | 5133 hCoV-19/England/CAMC-B09B38/2020\|EPI_ISL_672986\|20 | ···················· | ···················· | ···················· | ···················· | ···················· | ···················· |
| U | 5134 hCoV-19/England/CAMC-B09549/2020\|EPI_ISL_672988\|20 | ···················· | ···················· | ···················· | ···················· | ···················· | ···················· |
| U | 5135 hCoV-19/England/CAMC-B09EE4/2020\|EPI_ISL_672989\|20 | ···················· | ···················· | ···················· | ···················· | ···················· | ···················· |
| U | 5136 hCoV-19/England/CAMC-B09D7E/2020\|EPI_ISL_672990\|20 | ···················· | ···················· | ···················· | ···················· | ···················· | ···················· |
| U | 5137 hCoV-19/England/CAMC-B08DBB/2020\|EPI_ISL_672991\|20 | ···················· | ···················· | ···················· | ···················· | ···················· | ···················· |
| U | 5138 hCoV-19/England/CAMC-B08F6A/2020\|EPI_ISL_672993\|20 | ···················· | ···················· | ···················· | ···················· | ···················· | ···················· |
| U | 5139 hCoV-19/England/CAMC-B0A1CC/2020\|EPI_ISL_672994\|20 | ···················· | ···················· | ···················· | ···················· | ···················· | ···················· |
| U | 5140 hCoV-19/England/CAMC-B09C17/2020\|EPI_ISL_672995\|20 | ···················· | ···················· | ···················· | ···················· | ·····T·············· | ···················· |
| U | 5141 hCoV-19/England/CAMC-B096EC/2020\|EPI_ISL_672996\|20 | ···················· | ·A·················· | ···················· | ···················· | ···················· | ···················· |
| U | 5142 hCoV-19/England/CAMC-B08BA2/2020\|EPI_ISL_672997\|20 | ···················· | ···················· | ···················· | ···················· | ···················· | ···················· |
| U | 5143 hCoV-19/England/CAMC-B0A0FC/2020\|EPI_ISL_672998\|20 | ···················· | ···················· | ···················· | ···················· | ···················· | ···················· |
| U | 5144 hCoV-19/England/CAMC-B0A0ED/2020\|EPI_ISL_672999\|20 | ···················· | ···················· | ···················· | ···················· | ···················· | ···················· |
| U | 5145 hCoV-19/England/ALDP-B82C16/2020\|EPI_ISL_675200\|20 | ···················· | ···················· | ···················· | ···················· | ···················· | ···················· |
| U | 5146 hCoV-19/England/ALDP-B846BC/2020\|EPI_ISL_675202\|20 | ···················· | ···················· | ···················· | ···················· | ···················· | ···················· |
| U | 5147 hCoV-19/England/ALDP-B83A39/2020\|EPI_ISL_675203\|20 | ···················· | ···················· | ···················· | ···················· | ···················· | ···················· |
| U | 5148 hCoV-19/England/ALDP-B846CB/2020\|EPI_ISL_675291\|20 | ···················· | ···················· | ···················· | ···················· | ···················· | ···················· |
| U | 5149 hCoV-19/England/MILK-B8F2AE/2020\|EPI_ISL_675292\|20 | ···················· | ···················· | ···················· | ···················· | ···················· | ···················· |
| U | 5150 hCoV-19/England/ALDP-B85730/2020\|EPI_ISL_675293\|20 | ···················· | ···················· | ···················· | ···················· | ···················· | ···················· |
| U | 5151 hCoV-19/England/ALDP-B82CDA/2020\|EPI_ISL_675296\|20 | ···················· | ···················· | ···················· | ···················· | ···················· | ···················· |
| U | 5152 hCoV-19/England/ALDP-B82DB9/2020\|EPI_ISL_675297\|20 | ···················· | ···················· | ···················· | ···················· | ···················· | ···················· |
| U | 5153 hCoV-19/England/ALDP-B845BF/2020\|EPI_ISL_675298\|20 | ···················· | ···················· | ···················· | ···················· | ···················· | ···················· |
| U | 5154 hCoV-19/England/ALDP-B85897/2020\|EPI_ISL_675299\|20 | ···················· | ···················· | ···················· | ···················· | ···················· | ···················· |
| U | 5155 hCoV-19/Sweden/20-53499/2020\|EPI_ISL_676502\|2020-1 | ···················· | ···················· | ···················· | ···················· | ···················· | ···················· |
| U | 5156 hCoV-19/Sweden/20-53524/2020\|EPI_ISL_676503\|2020-1 | ···················· | ···················· | ···················· | ···················· | ···················· | ···················· |
| U | 5157 hCoV-19/Sweden/20-53525/2020\|EPI_ISL_676504\|2020-1 | ···················· | ···················· | ···················· | ···················· | ···················· | ···················· |
| U | 5158 hCoV-19/Sweden/20-53532/2020\|EPI_ISL_676505\|2020-1 | ···················· | ···················· | ···················· | ···················· | ···················· | ···················· |

| | | | | | | | | |
|---|---|---|---|---|---|---|---|---|
| U | 5159 hCoV-19/Sweden/20-53537/2020\|EPI_ISL_676506\|2020-0 | ···················· | ···················· | ···················· | ···················· | ···················· | ···················· | |
| U | 5160 hCoV-19/Sweden/20-53540/2020\|EPI_ISL_676507\|2020-1 | ···················· | ···················· | ···················· | ···················· | ···················· | ···················· | |
| U | 5161 hCoV-19/Sweden/20-53543/2020\|EPI_ISL_676508\|2020-1 | ···················· | ····A··············· | ···················· | ···················· | ···················· | ···················· | |
| U | 5162 hCoV-19/England/204881375/2020\|EPI_ISL_676511\|2020 | ···················· | ·A·················· | ···················· | ···················· | ···················· | ···················· | |
| U | 5163 hCoV-19/Sweden/20-08979/2020\|EPI_ISL_676512\|2020-0 | ···················· | ···················· | ···················· | ···················· | ···················· | ···················· | |
| U | 5164 hCoV-19/Sweden/20-22131/2020\|EPI_ISL_676513\|2020-0 | ···················· | ···················· | ···················· | ···················· | ···················· | ···················· | |
| U | 5165 hCoV-19/Sweden/20-52540/2020\|EPI_ISL_676514\|2020-0 | ···················· | ···················· | ···················· | ···················· | ···················· | ···················· | |
| U | 5166 hCoV-19/Sweden/20-53258/2020\|EPI_ISL_676515\|2020-1 | ···················· | ···················· | ···················· | ···················· | ···················· | ···················· | |
| U | 5167 hCoV-19/Sweden/20-08962/2020\|EPI_ISL_676519\|2020-0 | ···················· | ·A·················· | ···················· | ···················· | ···················· | ···················· | |
| U | 5168 hCoV-19/Sweden/20-08971/2020\|EPI_ISL_676520\|2020-0 | ···················· | ···················· | ···················· | ···················· | ···················· | ···················· | |
| U | 5169 hCoV-19/Sweden/20-21486/2020\|EPI_ISL_676521\|2020-0 | ···················· | ···················· | ···················· | ···················· | ···················· | ···················· | |
| U | 5170 hCoV-19/Sweden/20-21489/2020\|EPI_ISL_676522\|2020-0 | ···················· | ···················· | ···················· | ···················· | ···················· | ···················· | |
| U | 5171 hCoV-19/Sweden/20-21490/2020\|EPI_ISL_676523\|2020-0 | ···················· | ···················· | ···················· | ···················· | ···················· | ···················· | |
| U | 5172 hCoV-19/Sweden/20-21491/2020\|EPI_ISL_676524\|2020-0 | ···················· | ···················· | ···················· | ···················· | ···················· | ···················· | |
| U | 5173 hCoV-19/Sweden/20-21492/2020\|EPI_ISL_676525\|2020-0 | ···················· | ···················· | ···················· | ···················· | ···················· | ···················· | |
| U | 5174 hCoV-19/Sweden/20-21493/2020\|EPI_ISL_676526\|2020-0 | ···················· | ···················· | ···················· | ···················· | ···················· | ···················· | |
| U | 5175 hCoV-19/Sweden/20-21508/2020\|EPI_ISL_676527\|2020-0 | ···················· | ···················· | ···················· | ···················· | ···················· | ···················· | |
| U | 5176 hCoV-19/Sweden/20-21510/2020\|EPI_ISL_676528\|2020-0 | ···················· | ······A············· | ···················· | ···················· | ···················· | ···················· | |
| U | 5177 hCoV-19/Sweden/20-21511/2020\|EPI_ISL_676529\|2020-0 | ···················· | ···················· | ···················· | ···················· | ···················· | ···················· | |
| U | 5178 hCoV-19/Sweden/20-21513/2020\|EPI_ISL_676530\|2020-0 | ···················· | ···················· | ···················· | ···················· | ···················· | ···················· | |
| U | 5179 hCoV-19/Sweden/20-22276/2020\|EPI_ISL_676531\|2020-0 | ···················· | ···················· | ···················· | ···················· | ···················· | ···················· | |
| U | 5180 hCoV-19/Sweden/20-22277/2020\|EPI_ISL_676532\|2020-0 | ···················· | ···················· | ···················· | ···················· | ···················· | ···················· | |
| U | 5181 hCoV-19/Sweden/20-53027/2020\|EPI_ISL_676533\|2020-0 | ···················· | ···················· | ···················· | ···················· | ···················· | ···················· | |
| U | 5182 hCoV-19/Sweden/20-53067/2020\|EPI_ISL_676534\|2020-1 | ···················· | ···················· | ···················· | ···················· | ···················· | ···················· | |
| U | 5183 hCoV-19/USA/CA-IGI-0581/2020\|EPI_ISL_677835\|2020-0 | ···················· | ···················· | ···················· | ···················· | ···················· | ···················· | |
| U | 5184 hCoV-19/USA/CA-IGI-0585/2020\|EPI_ISL_677837\|2020-0 | ···················· | ···················· | ···················· | ···················· | ···················· | ···················· | |
| U | 5185 hCoV-19/USA/CA-IGI-0592/2020\|EPI_ISL_677838\|2020-0 | ···················· | ···················· | ···················· | ···················· | ···················· | ···················· | |
| U | 5186 hCoV-19/USA/CA-IGI-0595/2020\|EPI_ISL_677840\|2020-0 | ···················· | ···················· | ···················· | ···················· | ···················· | ···················· | |
| U | 5187 hCoV-19/USA/CA-IGI-0598/2020\|EPI_ISL_677841\|2020-0 | ···················· | ·A·················· | ···················· | ···················· | ···················· | ···················· | |
| U | 5188 hCoV-19/USA/CA-IGI-0602/2020\|EPI_ISL_677842\|2020-0 | ···················· | ···················· | ···················· | ···················· | ···················· | ···················· | |
| U | 5189 hCoV-19/USA/CA-IGI-0604/2020\|EPI_ISL_677843\|2020-0 | ···················· | ···················· | ···················· | ···················· | ···················· | ···················· | |
| U | 5190 hCoV-19/USA/CA-IGI-0608/2020\|EPI_ISL_677844\|2020-0 | ···················· | ···················· | ···················· | ···················· | ···················· | ···················· | |
| U | 5191 hCoV-19/USA/CA-IGI-0609/2020\|EPI_ISL_677845\|2020-0 | ···················· | ···················· | ···················· | ···················· | ···················· | ···················· | |
| U | 5192 hCoV-19/USA/CA-IGI-0613/2020\|EPI_ISL_677846\|2020-0 | ···················· | ·A·················· | ···················· | ···················· | ···················· | ···················· | |
| U | 5193 hCoV-19/USA/CA-IGI-0614/2020\|EPI_ISL_677847\|2020-0 | ···················· | ···················· | ···················· | ···················· | ···················· | ···················· | |
| U | 5194 hCoV-19/USA/CA-IGI-0618/2020\|EPI_ISL_677848\|2020-0 | ···················· | ···················· | ···················· | ···················· | ···················· | ···················· | |
| U | 5195 hCoV-19/USA/CA-IGI-0621/2020\|EPI_ISL_677849\|2020-0 | ···················· | ·A·················· | ···················· | ···················· | ···················· | ···················· | |
| U | 5196 hCoV-19/USA/CA-IGI-0622/2020\|EPI_ISL_677850\|2020-0 | ···················· | ···················· | ···················· | ···················· | ···················· | ···················· | |
| U | 5197 hCoV-19/USA/CA-IGI-0624/2020\|EPI_ISL_677851\|2020-0 | ···················· | ···················· | ···················· | ···················· | ···················· | ···················· | |
| U | 5198 hCoV-19/USA/CA-IGI-0625/2020\|EPI_ISL_677852\|2020-0 | ···················· | ···················· | ···················· | ···················· | ···················· | ···················· | |
| U | 5199 hCoV-19/USA/CA-IGI-0634/2020\|EPI_ISL_677853\|2020-0 | ···················· | ···················· | ···················· | ···················· | ···················· | ···················· | |
| U | 5200 hCoV-19/USA/CA-IGI-0635/2020\|EPI_ISL_677854\|2020-0 | ···················· | ···················· | ···················· | ···················· | ···················· | ···················· | |
| U | 5201 hCoV-19/USA/CA-IGI-0639/2020\|EPI_ISL_677855\|2020-0 | ···················· | ···················· | ···················· | ···················· | ···················· | ···················· | |
| U | 5202 hCoV-19/USA/CA-IGI-0641/2020\|EPI_ISL_677856\|2020-0 | ···················· | ···················· | ···················· | ···················· | ···················· | ···················· | |
| U | 5203 hCoV-19/USA/CA-IGI-0642/2020\|EPI_ISL_677857\|2020-0 | ···················· | ·A·················· | ···················· | ···················· | ···················· | ···················· | |
| U | 5204 hCoV-19/USA/CA-IGI-0647/2020\|EPI_ISL_677858\|2020-0 | ···················· | ···················· | ···················· | ···················· | ···················· | ···················· | |
| U | 5205 hCoV-19/USA/CA-IGI-0648/2020\|EPI_ISL_677859\|2020-0 | ···················· | ···················· | ···················· | ···················· | ···················· | ···················· | |
| U | 5206 hCoV-19/USA/CA-IGI-0652/2020\|EPI_ISL_677860\|2020-0 | ···················· | ···················· | ···················· | ···················· | ···················· | ···················· | |
| U | 5207 hCoV-19/USA/CA-IGI-0653/2020\|EPI_ISL_677861\|2020-0 | ···················· | ···················· | ···················· | ···················· | ···················· | ···················· | |
| U | 5208 hCoV-19/USA/CA-IGI-0656/2020\|EPI_ISL_677862\|2020-0 | ···················· | ···················· | ···················· | ···················· | ···················· | ···················· | |
| U | 5209 hCoV-19/USA/CA-IGI-0671/2020\|EPI_ISL_677864\|2020-0 | ···················· | ···················· | ···················· | ···················· | ···················· | ···················· | |
| U | 5210 hCoV-19/USA/CA-IGI-0672/2020\|EPI_ISL_677865\|2020-0 | ···················· | ····A··············· | ···················· | ···················· | ···················· | ···················· | |
| U | 5211 hCoV-19/Wales/PHWC-48ED69/2020\|EPI_ISL_681000\|2020 | ···················· | ···················· | ···················· | ···················· | ···················· | ···················· | |
| U | 5212 hCoV-19/Wales/PHWC-48ED87/2020\|EPI_ISL_681002\|2020 | ···················· | ···················· | ···················· | ···················· | ···················· | ···················· | |
| U | 5213 hCoV-19/Wales/PHWC-48ED96/2020\|EPI_ISL_681003\|2020 | ···················· | ···················· | ···················· | ···················· | ···················· | ···················· | |
| U | 5214 hCoV-19/Wales/PHWC-48EDA5/2020\|EPI_ISL_681004\|2020 | ···················· | ···················· | ···················· | ···················· | ···················· | ···················· | |
| U | 5215 hCoV-19/Wales/PHWC-48EDB4/2020\|EPI_ISL_681005\|2020 | ···················· | ···················· | ···················· | ···················· | ···················· | ···················· | |
| U | 5216 hCoV-19/Wales/PHWC-48EDC3/2020\|EPI_ISL_681006\|2020 | ···················· | ···················· | ···················· | ···················· | ···················· | ···················· | |
| U | 5217 hCoV-19/Wales/PHWC-48EDD2/2020\|EPI_ISL_681007\|2020 | ···················· | ···················· | ···················· | ···················· | ···················· | ···················· | |
| U | 5219 hCoV-19/Wales/PHWC-48EE0C/2020\|EPI_ISL_681009\|2020 | ···················· | ··········Y········· | ···················· | ···················· | ···················· | ···················· | |
| U | 5220 hCoV-19/Wales/PHWC-48EE1B/2020\|EPI_ISL_681010\|2020 | ···················· | ···················· | ···················· | ···················· | ···················· | ···················· | |
| U | 5221 hCoV-19/Wales/PHWC-48EE2A/2020\|EPI_ISL_681011\|2020 | ···················· | ···················· | ···················· | ···················· | ···················· | ···················· | |
| U | 5223 hCoV-19/Wales/PHWC-48EE66/2020\|EPI_ISL_681014\|2020 | ···················· | ···················· | ···················· | ···················· | ···················· | ···················· | |
| U | 5225 hCoV-19/Wales/PHWC-48EEA2/2020\|EPI_ISL_681017\|2020 | ···················· | ···················· | ···················· | ···················· | ···················· | ···················· | |
| U | 5226 hCoV-19/Wales/PHWC-48EEEE/2020\|EPI_ISL_681019\|2020 | ···················· | ···················· | ···················· | ···················· | ···················· | ···················· | |
| U | 5229 hCoV-19/Wales/PHWC-48EF72/2020\|EPI_ISL_681026\|2020 | ···················· | ···················· | ···················· | ···················· | ···················· | ···················· | |
| U | 5230 hCoV-19/Wales/PHWC-48EF90/2020\|EPI_ISL_681028\|2020 | ···················· | ···················· | ···················· | ···················· | ···················· | ·············NNN···· | 1 |
| U | 5232 hCoV-19/Wales/PHWC-48EFDC/2020\|EPI_ISL_681032\|2020 | ···················· | ···················· | ···················· | ···················· | ···················· | ···················· | |
| U | 5234 hCoV-19/Wales/PHWC-48EFFA/2020\|EPI_ISL_681034\|2020 | ···················· | ···················· | ···················· | ···················· | ···················· | ···················· | |
| U | 5235 hCoV-19/Wales/PHWC-48F005/2020\|EPI_ISL_681035\|2020 | ···················· | ···················· | ···················· | ···················· | ···················· | ···················· | |
| U | 5236 hCoV-19/Wales/PHWC-48F023/2020\|EPI_ISL_681036\|2020 | ···················· | ···················· | ···················· | ···················· | ···················· | ···················· | |
| U | 5237 hCoV-19/Wales/PHWC-48F032/2020\|EPI_ISL_681037\|2020 | ···················· | ···················· | ···················· | ···················· | ···················· | ···················· | |
| U | 5239 hCoV-19/Wales/PHWC-48F050/2020\|EPI_ISL_681039\|2020 | ···················· | ···················· | ···················· | ···················· | ···················· | ···················· | |
| U | 5240 hCoV-19/Wales/PHWC-48F09C/2020\|EPI_ISL_681040\|2020 | ···················· | ···················· | ···················· | ···················· | ···················· | ···················· | |
| U | 5241 hCoV-19/Wales/PHWC-48F0BA/2020\|EPI_ISL_681042\|2020 | ···················· | ···················· | ···················· | ···················· | ···················· | ···················· | |
| U | 5242 hCoV-19/Wales/PHWC-48F0D8/2020\|EPI_ISL_681043\|2020 | ···················· | ···················· | ···················· | ···················· | ···················· | ···················· | |
| U | 5243 hCoV-19/Bahrain/340685804/2020\|EPI_ISL_682300\|2020 | ···················· | ···················· | ···················· | ···················· | ···················· | ···················· | |
| U | 5244 hCoV-19/Bahrain/260104364/2020\|EPI_ISL_682301\|2020 | ···················· | ···················· | ···················· | ···················· | ···················· | ···················· | |
| U | 5245 hCoV-19/Bahrain/340958054/2020\|EPI_ISL_682302\|2020 | ···················· | ···················· | ···················· | ···················· | ···················· | ···················· | |
| U | 5246 hCoV-19/Bahrain/920407807/2020\|EPI_ISL_682303\|2020 | ···················· | ···················· | ···················· | ···················· | ···················· | ···················· | |

| | | | | | | | | |
|---|---|---|---|---|---|---|---|---|
| U | 5247 | hCoV-19/Bahrain/920457147/2020\|EPI_ISL_682304\|2020 | ···················· | ···················· | ···················· | ···················· | ···················· | ···················· |
| U | 5248 | hCoV-19/Bahrain/920456050/2020\|EPI_ISL_682305\|2020 | ···················· | ···················· | ···················· | ···················· | ···················· | ···················· |
| U | 5249 | hCoV-19/Bahrain/920455400/2020\|EPI_ISL_682306\|2020 | ···················· | ···················· | ···················· | ···················· | ···················· | ···················· |
| U | 5250 | hCoV-19/Bahrain/920453170/2020\|EPI_ISL_682307\|2020 | ···················· | ···················· | ···················· | ···················· | ···················· | ···················· |
| U | 5251 | hCoV-19/Bahrain/920453107/2020\|EPI_ISL_682308\|2020 | ···················· | ···················· | ···················· | ···················· | ···················· | ···················· |
| U | 5252 | hCoV-19/Bahrain/920453092/2020\|EPI_ISL_682309\|2020 | ···················· | ···················· | ···················· | ···················· | ···················· | ···················· |
| U | 5253 | hCoV-19/Bahrain/600022987/2020\|EPI_ISL_682310\|2020 | ···················· | ···················· | ···················· | ···················· | ···················· | ···················· |
| U | 5254 | hCoV-19/Bahrain/340861249/2020\|EPI_ISL_682311\|2020 | ···················· | ···················· | ···················· | ···················· | ···················· | ···················· |
| U | 5255 | hCoV-19/Bahrain/340854790/2020\|EPI_ISL_682312\|2020 | ···················· | ···················· | ···················· | ···················· | ···················· | ···················· |
| U | 5256 | hCoV-19/Bahrain/340854759/2020\|EPI_ISL_682313\|2020 | ···················· | ···················· | ···················· | ···················· | ···················· | ···················· |
| U | 5257 | hCoV-19/Bahrain/340846472/2020\|EPI_ISL_682314\|2020 | ···················· | ···················· | ···················· | ···················· | ···················· | ···················· |
| U | 5258 | hCoV-19/Bahrain/250182773/2020\|EPI_ISL_682315\|2020 | ···················· | ···················· | ···················· | ···················· | ···················· | ···················· |
| U | 5259 | hCoV-19/Bahrain/250182774/2020\|EPI_ISL_682316\|2020 | ···················· | ···················· | ···················· | ···················· | ···················· | ···················· |
| U | 5260 | hCoV-19/Bahrain/341015978/2020\|EPI_ISL_682317\|2020 | ···················· | ···················· | ···················· | ···················· | ···················· | ···················· |
| U | 5261 | hCoV-19/Bahrain/341017087/2020\|EPI_ISL_682318\|2020 | ···················· | ···················· | ···················· | ···················· | ···················· | ···················· |
| U | 5262 | hCoV-19/Bahrain/341036667/2020\|EPI_ISL_682319\|2020 | ···················· | ·A·················· | ···················· | ···················· | ···················· | ··A················· |
| U | 5263 | hCoV-19/Bahrain/341038022/2020\|EPI_ISL_682320\|2020 | ···················· | ···A················ | ···················· | ···················· | ···················· | ··A················· |
| U | 5264 | hCoV-19/Bahrain/341050336/2020\|EPI_ISL_682321\|2020 | ···················· | ···················· | ···················· | ···················· | ···················· | ···················· |
| U | 5265 | hCoV-19/Bahrain/341067576/2020\|EPI_ISL_682322\|2020 | ···················· | ···················· | ···················· | ···················· | ···················· | ···················· |
| U | 5266 | hCoV-19/South Africa/UFS-VIRO-NGS-10/2020\|EPI_ISL_ | ···················· | ···················· | ···················· | ···················· | ···················· | ···················· |
| U | 5267 | hCoV-19/South Africa/UFS-VIRO-NGS-11/2020\|EPI_ISL_ | ···················· | ···················· | ···················· | ···················· | ···················· | ···················· |
| U | 5268 | hCoV-19/South Africa/UFS-VIRO-NGS-12/2020\|EPI_ISL_ | ···················· | ·A·················· | ···················· | ···················· | ···················· | ···················· |
| U | 5269 | hCoV-19/South Africa/UFS-VIRO-NGS-14/2020\|EPI_ISL_ | ···················· | ·A·················· | ···················· | ···················· | ···················· | ···················· |
| U | 5270 | hCoV-19/South Africa/UFS-VIRO-NGS-17/2020\|EPI_ISL_ | ···················· | ···················· | ···················· | ···················· | ···················· | ···················· |
| U | 5271 | hCoV-19/South Africa/UFS-VIRO-NGS-18/2020\|EPI_ISL_ | ···················· | ···················· | ···················· | ···················· | ···················· | ···················· |
| U | 5272 | hCoV-19/South Africa/UFS-VIRO-NGS-20/2020\|EPI_ISL_ | ···················· | ···················· | ···················· | ···················· | ···················· | ···················· |
| U | 5273 | hCoV-19/South Africa/UFS-VIRO-NGS-22/2020\|EPI_ISL_ | ···················· | ·A·················· | ···················· | ···················· | ···················· | ···················· |
| U | 5274 | hCoV-19/South Africa/UFS-VIRO-NGS-25/2020\|EPI_ISL_ | ···················· | ···················· | ···················· | ···················· | ···················· | ···················· |
| U | 5275 | hCoV-19/South Africa/UFS-VIRO-NGS-27/2020\|EPI_ISL_ | ···················· | ·A·················· | ···················· | ···················· | ···················· | ···················· |
| U | 5276 | hCoV-19/South Africa/UFS-VIRO-NGS-28/2020\|EPI_ISL_ | ···················· | ···················· | ···················· | ···················· | ···················· | ···················· |
| U | 5277 | hCoV-19/South Africa/UFS-VIRO-NGS-29/2020\|EPI_ISL_ | ···················· | ·A·················· | ···················· | ···················· | ···················· | ···················· |
| U | 5278 | hCoV-19/South Africa/UFS-VIRO-NGS-30/2020\|EPI_ISL_ | ···················· | ·A·················· | ···················· | ···················· | ···················· | ···················· |
| U | 5279 | hCoV-19/South Africa/UFS-VIRO-NGS-31/2020\|EPI_ISL_ | ···················· | ·A·················· | ···················· | ···················· | ···················· | ···················· |
| U | 5280 | hCoV-19/South Africa/UFS-VIRO-NGS-34/2020\|EPI_ISL_ | ···················· | ···················· | ···················· | ···················· | ···················· | ···················· |
| U | 5281 | hCoV-19/South Africa/UFS-VIRO-NGS-35/2020\|EPI_ISL_ | ···················· | ·A·················· | ···················· | ···················· | ···················· | ···················· |
| U | 5282 | hCoV-19/South Africa/UFS-VIRO-NGS-39/2020\|EPI_ISL_ | ···················· | ···················· | ···················· | ···················· | ···················· | ···················· |
| U | 5283 | hCoV-19/South Africa/UFS-VIRO-NGS-41/2020\|EPI_ISL_ | ···················· | ·A·················· | ···················· | ···················· | ···················· | ···················· |
| U | 5284 | hCoV-19/South Africa/UFS-VIRO-NGS-7/2020\|EPI_ISL_6 | ···················· | ···················· | ···················· | ···················· | ···················· | ···················· |
| U | 5285 | hCoV-19/South Africa/UFS-VIRO-NGS-8/2020\|EPI_ISL_6 | ···················· | ···················· | ···················· | ···················· | ···················· | ···················· |
| U | 5286 | hCoV-19/Denmark/DCGC-12172/2020\|EPI_ISL_682353\|202 | ···················· | ···················· | ···················· | ···················· | ···················· | ···················· |
| U | 5287 | hCoV-19/Denmark/DCGC-12173/2020\|EPI_ISL_682354\|202 | ···················· | ·A·················· | ···················· | ···················· | ···················· | ···················· |
| U | 5288 | hCoV-19/Denmark/DCGC-12176/2020\|EPI_ISL_682357\|202 | ···················· | ·A·················· | ···················· | ···················· | ···················· | ···················· |
| U | 5289 | hCoV-19/Denmark/DCGC-12177/2020\|EPI_ISL_682358\|202 | ···················· | ···················· | ···················· | ···················· | ···················· | ···················· |
| U | 5290 | hCoV-19/Denmark/DCGC-12180/2020\|EPI_ISL_682361\|202 | ···················· | ···················· | ···················· | ···················· | ···················· | ···················· |
| U | 5291 | hCoV-19/Denmark/DCGC-12181/2020\|EPI_ISL_682362\|202 | ···················· | ···················· | ···················· | ···················· | ···················· | ···················· |
| U | 5292 | hCoV-19/Denmark/DCGC-12182/2020\|EPI_ISL_682363\|202 | ···················· | ···················· | ···················· | ···················· | ···················· | ···················· |
| U | 5293 | hCoV-19/Denmark/DCGC-12183/2020\|EPI_ISL_682364\|202 | ···················· | ···················· | ···················· | ···················· | ···················· | ···················· |
| U | 5294 | hCoV-19/Denmark/DCGC-12184/2020\|EPI_ISL_682365\|202 | ···················· | ···················· | ···················· | ···················· | ···················· | ···················· |
| U | 5295 | hCoV-19/Denmark/DCGC-12185/2020\|EPI_ISL_682366\|202 | ···················· | ·A·················· | ···················· | ···················· | ···················· | ··A················· |
| U | 5296 | hCoV-19/Denmark/DCGC-12186/2020\|EPI_ISL_682367\|202 | ···················· | ···················· | ···················· | ···················· | ···················· | ···················· |
| U | 5297 | hCoV-19/Denmark/DCGC-12188/2020\|EPI_ISL_682369\|202 | ···················· | ·A·················· | ···················· | ···················· | ···················· | ··A················· |
| U | 5298 | hCoV-19/Denmark/DCGC-12189/2020\|EPI_ISL_682370\|202 | ···················· | ···················· | ···················· | ···················· | ···················· | ···················· |
| U | 5299 | hCoV-19/Denmark/DCGC-12190/2020\|EPI_ISL_682371\|202 | ···················· | ···················· | ···················· | ···················· | ···················· | ···················· |
| U | 5300 | hCoV-19/Denmark/DCGC-12191/2020\|EPI_ISL_682372\|202 | ···················· | ···················· | ···················· | ···················· | ···················· | ···················· |
| U | 5301 | hCoV-19/Denmark/DCGC-12193/2020\|EPI_ISL_682374\|202 | ···················· | ···················· | ···················· | ···················· | ···················· | ···················· |
| U | 5302 | hCoV-19/Denmark/DCGC-12194/2020\|EPI_ISL_682375\|202 | ···················· | ···················· | ···················· | ···················· | ···················· | ···················· |
| U | 5303 | hCoV-19/Denmark/DCGC-12220/2020\|EPI_ISL_682401\|202 | ···················· | ···················· | ···················· | ···················· | ···················· | ···················· |
| U | 5325 | hCoV-19/USA/MN-MDH-2049/2020\|EPI_ISL_683678\|2020-0 | ···················· | ···················· | ···················· | ···················· | ···················· | ···················· |
| U | 5326 | hCoV-19/USA/MN-MDH-2050/2020\|EPI_ISL_683679\|2020-0 | ···················· | ···················· | ···················· | ···················· | ···················· | ···················· |
| U | 5327 | hCoV-19/USA/MN-MDH-2051/2020\|EPI_ISL_683680\|2020-0 | ···················· | ···················· | ···················· | ···················· | ···················· | ···················· |
| U | 5328 | hCoV-19/USA/MN-MDH-2052/2020\|EPI_ISL_683681\|2020-0 | ···················· | ···················· | ···················· | ···················· | ···················· | ···················· |
| U | 5329 | hCoV-19/USA/MN-MDH-2053/2020\|EPI_ISL_683682\|2020-0 | ···················· | ·A·················· | ···················· | ···················· | ···················· | ···················· |
| U | 5330 | hCoV-19/USA/MN-MDH-2054/2020\|EPI_ISL_683683\|2020-0 | ···················· | ···················· | ···················· | ···················· | ···················· | ···················· |
| U | 5331 | hCoV-19/USA/MN-MDH-2055/2020\|EPI_ISL_683684\|2020-0 | ···················· | ···················· | ···················· | ···················· | ···················· | ···················· |
| U | 5332 | hCoV-19/USA/MN-MDH-2056/2020\|EPI_ISL_683685\|2020-0 | ···················· | ···················· | ···················· | ···················· | ···················· | ··R················· |
| U | 5333 | hCoV-19/USA/MN-MDH-2057/2020\|EPI_ISL_683686\|2020-0 | ···················· | ···················· | ···················· | ···················· | ···················· | ···················· |
| U | 5334 | hCoV-19/USA/MN-MDH-2058/2020\|EPI_ISL_683687\|2020-0 | ···················· | ···················· | ···················· | ···················· | ···················· | ···················· |
| U | 5335 | hCoV-19/USA/MN-MDH-2059/2020\|EPI_ISL_683688\|2020-0 | ···················· | ···················· | ···················· | ···················· | ···················· | ···················· |
| U | 5336 | hCoV-19/USA/MN-MDH-2060/2020\|EPI_ISL_683689\|2020-1 | ···················· | ·A·················· | ···················· | ···················· | ···················· | ···················· |
| U | 5337 | hCoV-19/USA/MN-MDH-2061/2020\|EPI_ISL_683690\|2020-1 | ···················· | ···················· | ···················· | ···················· | ···················· | ···················· |
| U | 5338 | hCoV-19/USA/MN-MDH-2062/2020\|EPI_ISL_683691\|2020-1 | ···················· | ·A·················· | ···················· | ···················· | ···················· | ···················· |
| U | 5339 | hCoV-19/USA/MN-MDH-2063/2020\|EPI_ISL_683692\|2020-1 | ···················· | ·A·················· | ···················· | ···················· | ···················· | ···················· |
| U | 5340 | hCoV-19/USA/MN-MDH-2064/2020\|EPI_ISL_683693\|2020-1 | ···················· | ·A·················· | ···················· | ···················· | ···················· | ···················· |
| U | 5341 | hCoV-19/USA/MN-MDH-2065/2020\|EPI_ISL_683694\|2020-1 | ···················· | ·A·················· | ···················· | ···················· | ···················· | ···················· |
| U | 5342 | hCoV-19/USA/MN-MDH-2066/2020\|EPI_ISL_683695\|2020-1 | ···················· | ···················· | ···················· | ···················· | ···················· | ···················· |
| U | 5343 | hCoV-19/USA/MN-MDH-2067/2020\|EPI_ISL_683696\|2020-1 | ···················· | ·A·················· | ···················· | ···················· | ···················· | ···················· |
| U | 5344 | hCoV-19/USA/MN-MDH-2068/2020\|EPI_ISL_683697\|2020-1 | ···················· | ···················· | ···················· | ···················· | ···················· | ···················· |
| U | 5345 | hCoV-19/USA/MN-MDH-2069/2020\|EPI_ISL_683698\|2020-1 | ···················· | ·A·················· | ···················· | ···················· | ···················· | ···················· |
| U | 5346 | hCoV-19/USA/MN-MDH-2070/2020\|EPI_ISL_683699\|2020-1 | ···················· | ·A·················· | ···················· | ···················· | ···················· | ···················· |
| U | 5347 | hCoV-19/USA/MN-MDH-2071/2020\|EPI_ISL_683700\|2020-1 | ···················· | ·A·················· | ···················· | ···················· | ···················· | ···················· |

| | | | | | | | |
|---|---|---|---|---|---|---|---|
| U | 5348 hCoV-19/USA/MN-MDH-2072/2020\|EPI_ISL_683701\|2020-1 | ···················· | ···················· | ···················· | ···················· | ···················· | ···················· |
| U | 5349 hCoV-19/USA/MN-MDH-2073/2020\|EPI_ISL_683702\|2020-1 | ···················· | ·A·················· | ···················· | ···················· | ···················· | ···················· |
| U | 5350 hCoV-19/USA/MN-MDH-2074/2020\|EPI_ISL_683703\|2020-1 | ···················· | ·A·················· | ···················· | ···················· | ···················· | ···················· |
| U | 5351 hCoV-19/USA/MN-MDH-2075/2020\|EPI_ISL_683704\|2020-1 | ···················· | ·A·················· | ···················· | ···················· | ···················· | ···················· |
| U | 5352 hCoV-19/USA/MN-MDH-2076/2020\|EPI_ISL_683705\|2020-1 | ···················· | ·A·················· | ···················· | ···················· | ···················· | ···················· |
| U | 5353 hCoV-19/USA/MN-MDH-2077/2020\|EPI_ISL_683706\|2020-1 | ···················· | ·A·················· | ···················· | ···················· | ···················· | ···················· |
| U | 5354 hCoV-19/USA/MN-MDH-2078/2020\|EPI_ISL_683707\|2020-1 | ···················· | ·A·················· | ···················· | ···················· | ···················· | ···················· |
| U | 5355 hCoV-19/USA/MN-MDH-2079/2020\|EPI_ISL_683708\|2020-1 | ···················· | ·A·················· | ···················· | ···················· | ···················· | ···················· |
| U | 5356 hCoV-19/USA/MN-MDH-2080/2020\|EPI_ISL_683709\|2020-1 | ···················· | ·A·················· | ···················· | ···················· | ···················· | ···················· |
| U | 5357 hCoV-19/USA/MN-MDH-2081/2020\|EPI_ISL_683710\|2020-1 | ···················· | ·A·················· | ···················· | ···················· | ···················· | ···················· |
| U | 5358 hCoV-19/USA/MN-MDH-2082/2020\|EPI_ISL_683711\|2020-1 | ···················· | ·A·················· | ···················· | ···················· | ···················· | ···················· |
| U | 5359 hCoV-19/USA/MN-MDH-2083/2020\|EPI_ISL_683712\|2020-1 | ···················· | ·A·················· | ···················· | ···················· | ···················· | ···················· |
| U | 5360 hCoV-19/USA/MN-MDH-2084/2020\|EPI_ISL_683713\|2020-1 | ···················· | ·A·················· | ···················· | ···················· | ···················· | ···················· |
| U | 5361 hCoV-19/USA/MN-MDH-2085/2020\|EPI_ISL_683714\|2020-1 | ···················· | ···················· | ···················· | ···················· | ···················· | ···················· |
| U | 5362 hCoV-19/USA/MN-MDH-2086/2020\|EPI_ISL_683715\|2020-1 | ···················· | ···················· | ···················· | ···················· | ···················· | ···················· |
| U | 5363 hCoV-19/USA/MN-MDH-2087/2020\|EPI_ISL_683716\|2020-1 | ···················· | ···················· | ···················· | ···················· | ···················· | ···················· |
| U | 5364 hCoV-19/USA/MN-MDH-2088/2020\|EPI_ISL_683717\|2020-1 | ···················· | ···················· | ···················· | ···················· | ···················· | ···················· |
| U | 5365 hCoV-19/USA/MN-MDH-2089/2020\|EPI_ISL_683718\|2020-1 | ···················· | ···················· | ···················· | ···················· | ···················· | ···················· |
| U | 5366 hCoV-19/USA/MN-MDH-2090/2020\|EPI_ISL_683719\|2020-1 | ···················· | ···················· | ···················· | ···················· | ···················· | ···················· |
| U | 5367 hCoV-19/USA/MN-MDH-2091/2020\|EPI_ISL_683720\|2020-1 | ···················· | ·A·················· | ···················· | ···················· | ···················· | ···················· |
| U | 5368 hCoV-19/USA/MN-MDH-2092/2020\|EPI_ISL_683721\|2020-1 | ···················· | ···················· | ···················· | ···················· | ···················· | ···················· |
| U | 5369 hCoV-19/USA/MN-MDH-2093/2020\|EPI_ISL_683722\|2020-1 | ···················· | ···················· | ···················· | ···················· | ···················· | ···················· |
| U | 5370 hCoV-19/USA/MN-MDH-2094/2020\|EPI_ISL_683723\|2020-1 | ···················· | ···················· | ···················· | ···················· | ···················· | ···················· |
| U | 5371 hCoV-19/USA/MN-MDH-2095/2020\|EPI_ISL_683724\|2020-1 | ···················· | ···················· | ···················· | ···················· | ···················· | ···················· |
| U | 5372 hCoV-19/USA/MN-MDH-2096/2020\|EPI_ISL_683725\|2020-1 | ···················· | ···················· | ···················· | ···················· | ···················· | ···················· |
| U | 5373 hCoV-19/USA/MN-MDH-2097/2020\|EPI_ISL_683726\|2020-1 | ···················· | ···················· | ···················· | ···················· | ···················· | ···················· |
| U | 5374 hCoV-19/USA/MN-MDH-2098/2020\|EPI_ISL_683727\|2020-1 | ···················· | ···················· | ···················· | ···················· | ···················· | ···················· |
| U | 5375 hCoV-19/USA/MN-MDH-2099/2020\|EPI_ISL_683728\|2020-1 | ···················· | ···················· | ···················· | ···················· | ···················· | ···················· |
| U | 5376 hCoV-19/USA/MN-MDH-2100/2020\|EPI_ISL_683729\|2020-1 | ···················· | ···················· | ···················· | ···················· | ···················· | ···················· |
| U | 5377 hCoV-19/England/MILK-B95E29/2020\|EPI_ISL_675100\|20 | ···················· | ···················· | ···················· | ···················· | ···················· | ···················· |
| U | 5378 hCoV-19/England/MILK-B97294/2020\|EPI_ISL_675101\|20 | ···················· | ···················· | ···················· | ···················· | ···················· | ···················· |
| U | 5379 hCoV-19/England/MILK-B96CD3/2020\|EPI_ISL_675103\|20 | ···················· | ···················· | ···················· | ···················· | ···················· | ···················· |
| U | 5380 hCoV-19/England/CAMC-B98169/2020\|EPI_ISL_675104\|20 | ···················· | ···················· | ···················· | ···················· | ···················· | ···················· |
| U | 5381 hCoV-19/England/MILK-B974CB/2020\|EPI_ISL_675105\|20 | ···················· | ···················· | ···················· | ···················· | ·····T·············· | ···················· |
| U | 5382 hCoV-19/England/MILK-B97452/2020\|EPI_ISL_675106\|20 | ···················· | ···················· | ···················· | ···················· | ···················· | ···················· |
| U | 5383 hCoV-19/England/CAMC-B986E2/2020\|EPI_ISL_675107\|20 | ···················· | ·A·················· | ···················· | ···················· | ···················· | ··A················· |
| U | 5384 hCoV-19/England/MILK-B96BF4/2020\|EPI_ISL_675108\|20 | ···················· | ···················· | ···················· | ···················· | ···················· | ···················· |
| U | 5385 hCoV-19/England/MILK-B972D0/2020\|EPI_ISL_675109\|20 | ···················· | ···················· | ···················· | ···················· | ···················· | ···················· |
| U | 5386 hCoV-19/England/CAMC-B981A5/2020\|EPI_ISL_675110\|20 | ···················· | ···················· | ···················· | ···················· | ···················· | ···················· |
| U | 5387 hCoV-19/England/MILK-B96ECE/2020\|EPI_ISL_675111\|20 | ···················· | ···················· | ···················· | ···················· | ···················· | ···················· |
| U | 5388 hCoV-19/England/MILK-B96D0D/2020\|EPI_ISL_675113\|20 | ···················· | ···················· | ···················· | ···················· | ···················· | ···················· |
| U | 5389 hCoV-19/England/MILK-B96E0A/2020\|EPI_ISL_675114\|20 | ···················· | ···················· | ···················· | ···················· | ···················· | ···················· |
| U | 5390 hCoV-19/England/MILK-B95AF8/2020\|EPI_ISL_675115\|20 | ···················· | ···················· | ···················· | ···················· | ···················· | ···················· |
| U | 5391 hCoV-19/England/MILK-B96B9A/2020\|EPI_ISL_675116\|20 | ···················· | ···················· | ···················· | ···················· | ···················· | ···················· |
| U | 5392 hCoV-19/England/CAMC-B98363/2020\|EPI_ISL_675117\|20 | ···················· | ···················· | ···················· | ···················· | ···················· | ···················· |
| U | 5393 hCoV-19/England/MILK-B96514/2020\|EPI_ISL_675119\|20 | ···················· | ···················· | ···················· | ···················· | ···················· | ···················· |
| U | 5394 hCoV-19/England/CAMC-B9804E/2020\|EPI_ISL_675120\|20 | ···················· | ···················· | ···················· | ···················· | ···················· | ···················· |
| U | 5395 hCoV-19/England/MILK-B959A0/2020\|EPI_ISL_675121\|20 | ···················· | ···················· | ···················· | ···················· | ···················· | ···················· |
| U | 5396 hCoV-19/England/MILK-B95973/2020\|EPI_ISL_675122\|20 | ·T·················· | ···················· | ···················· | ···················· | ···················· | ···················· |
| U | 5397 hCoV-19/England/MILK-B9755F/2020\|EPI_ISL_675124\|20 | ···················· | ···················· | ···················· | ···················· | ···················· | ···················· |
| U | 5398 hCoV-19/England/MILK-B96C79/2020\|EPI_ISL_675125\|20 | ···················· | ···················· | ···················· | ···················· | ···················· | ···················· |
| U | 5399 hCoV-19/England/CAMC-B984AC/2020\|EPI_ISL_675126\|20 | ···················· | ···················· | ···················· | ···················· | ···················· | ···················· |
| U | 5400 hCoV-19/England/MILK-B95919/2020\|EPI_ISL_675127\|20 | ···················· | ···················· | ···················· | ···················· | ···················· | ···················· |
| U | 5401 hCoV-19/England/ALDP-B84160/2020\|EPI_ISL_675130\|20 | ···················· | ···················· | ···················· | ···················· | ···················· | ···················· |
| U | 5402 hCoV-19/England/MILK-B8F1A1/2020\|EPI_ISL_675131\|20 | ···················· | ···················· | ···················· | ···················· | ···················· | ···················· |
| U | 5403 hCoV-19/England/MILK-B8F3E7/2020\|EPI_ISL_675132\|20 | ···················· | ···················· | ···················· | ···················· | ···················· | ···················· |
| U | 5404 hCoV-19/England/ALDP-B8477D/2020\|EPI_ISL_675133\|20 | ···················· | ···················· | ···················· | ···················· | ···················· | ···················· |
| U | 5405 hCoV-19/England/MILK-B8F332/2020\|EPI_ISL_675134\|20 | ···················· | ···················· | ···················· | ···················· | ···················· | ···················· |
| U | 5406 hCoV-19/England/ALDP-B83A75/2020\|EPI_ISL_675135\|20 | ···················· | ···················· | ···················· | ···················· | ···················· | ···················· |
| U | 5407 hCoV-19/England/ALDP-B84546/2020\|EPI_ISL_675136\|20 | ···················· | ···················· | ···················· | ···················· | ···················· | ···················· |
| U | 5408 hCoV-19/England/ALDP-B82D6E/2020\|EPI_ISL_675137\|20 | ···················· | ···················· | ···················· | ···················· | ···················· | ···················· |
| U | 5409 hCoV-19/England/ALDP-B83FEF/2020\|EPI_ISL_675138\|20 | ···················· | ···················· | ···················· | ···················· | ···················· | ···················· |
| U | 5410 hCoV-19/England/ALDP-B8577C/2020\|EPI_ISL_675139\|20 | ···················· | ···················· | ···················· | ···················· | ···················· | ···················· |
| U | 5411 hCoV-19/England/ALDP-B82CBC/2020\|EPI_ISL_675140\|20 | ···················· | ···················· | ···················· | ···················· | ···················· | ···················· |
| U | 5412 hCoV-19/England/ALDP-B8392D/2020\|EPI_ISL_675141\|20 | ···················· | ···················· | ···················· | ···················· | ···················· | ···················· |
| U | 5413 hCoV-19/England/ALDP-B83C33/2020\|EPI_ISL_675142\|20 | ···················· | ···················· | ···················· | ···················· | ···················· | ···················· |
| U | 5414 hCoV-19/England/ALDP-B8388A/2020\|EPI_ISL_675143\|20 | ···················· | ···················· | ···················· | ···················· | ···················· | ···················· |
| U | 5415 hCoV-19/England/ALDP-B84476/2020\|EPI_ISL_675144\|20 | ···················· | ···················· | ···················· | ···················· | ···················· | ···················· |
| U | 5416 hCoV-19/England/MILK-B8F39C/2020\|EPI_ISL_675145\|20 | ···················· | ···················· | ···················· | ···················· | ···················· | ···················· |
| U | 5417 hCoV-19/England/ALDP-B8578B/2020\|EPI_ISL_675146\|20 | ···················· | ···················· | ···················· | ···················· | ···················· | ···················· |
| U | 5418 hCoV-19/England/ALDP-B83C42/2020\|EPI_ISL_675147\|20 | ···················· | ···················· | ···················· | ···················· | ···················· | ···················· |
| U | 5419 hCoV-19/England/ALDP-B836CC/2020\|EPI_ISL_675148\|20 | ···················· | ···················· | ···················· | ···················· | ···················· | ···················· |
| U | 5420 hCoV-19/England/ALDP-B84555/2020\|EPI_ISL_675150\|20 | ···················· | ···················· | ···················· | ···················· | ···················· | ···················· |
| U | 5421 hCoV-19/England/ALDP-B83CE8/2020\|EPI_ISL_675151\|20 | ···················· | ···················· | ···················· | ···················· | ···················· | ···················· |
| U | 5422 hCoV-19/England/ALDP-B83C51/2020\|EPI_ISL_675152\|20 | ···················· | ···················· | ···················· | ···················· | ···················· | ···················· |
| U | 5423 hCoV-19/England/MILK-B8F01D/2020\|EPI_ISL_675153\|20 | ···················· | ···················· | ···················· | ···················· | ···················· | ···················· |
| U | 5424 hCoV-19/England/ALDP-B83E97/2020\|EPI_ISL_675154\|20 | ···················· | ···················· | ···················· | ···················· | ···················· | ···················· |
| U | 5425 hCoV-19/England/ALDP-B8579A/2020\|EPI_ISL_675155\|20 | ···················· | ···················· | ···················· | ···················· | ···················· | ···················· |
| U | 5426 hCoV-19/England/ALDP-B857D6/2020\|EPI_ISL_675156\|20 | ···················· | ···················· | ···················· | ···················· | ···················· | ···················· |
| U | 5427 hCoV-19/England/ALDP-B841F7/2020\|EPI_ISL_675157\|20 | ···················· | ···················· | ···················· | ···················· | ···················· | ···················· |

| | | | | | | | |
|---|---|---|---|---|---|---|---|
| U | 5428 hCoV-19/England/ALDP-B84582/2020\|EPI_ISL_675158\|20 | .................... | .................... | ..................... | .................... | .................... | .................... |
| U | 5429 hCoV-19/England/ALDP-B83A2A/2020\|EPI_ISL_675159\|20 | .................... | .................... | ..................... | .................... | .................... | .................... |
| U | 5430 hCoV-19/England/MILK-B8F3BA/2020\|EPI_ISL_675160\|20 | .................... | .................... | ..................... | .................... | ......T............. | .................... |
| U | 5431 hCoV-19/England/ALDP-B849A4/2020\|EPI_ISL_675161\|20 | .................... | .................... | ..................... | .................... | .................... | .................... |
| U | 5432 hCoV-19/England/ALDP-B82CE9/2020\|EPI_ISL_675163\|20 | .................... | .................... | ..................... | .................... | .................... | .................... |
| U | 5433 hCoV-19/England/ALDP-B82A0D/2020\|EPI_ISL_675164\|20 | .................... | .................... | ..................... | .................... | .................... | .................... |
| U | 5434 hCoV-19/England/MILK-B8EF7B/2020\|EPI_ISL_675165\|20 | .................... | .................... | ..................... | .................... | .................... | .................... |
| U | 5435 hCoV-19/England/ALDP-B835A1/2020\|EPI_ISL_675166\|20 | .................... | .................... | ..................... | .................... | .................... | .................... |
| U | 5436 hCoV-19/England/ALDP-B836F9/2020\|EPI_ISL_675167\|20 | .................... | .................... | ..................... | .................... | .................... | .................... |
| U | 5437 hCoV-19/England/ALDP-B84591/2020\|EPI_ISL_675168\|20 | .................... | .................... | ..................... | .................... | .................... | .................... |
| U | 5438 hCoV-19/England/ALDP-B82F0E/2020\|EPI_ISL_675169\|20 | .................... | .................... | ..................... | .................... | .................... | .................... |
| U | 5439 hCoV-19/Scotland/QEUH-B3187C/2020\|EPI_ISL_676420\|2 | .................... | .................... | ..................... | .................... | .................... | .................... |
| U | 5440 hCoV-19/Scotland/QEUH-B31E01/2020\|EPI_ISL_676421\|2 | .................... | .................... | ..................... | .................... | ......T............. | .................... |
| U | 5441 hCoV-19/Scotland/QEUH-B3179D/2020\|EPI_ISL_676422\|2 | .................... | .................... | ..................... | .................... | .................... | .................... |
| U | 5442 hCoV-19/Scotland/QEUH-B3178E/2020\|EPI_ISL_676425\|2 | .................... | .................... | ..................... | .................... | .................... | .................... |
| U | 5443 hCoV-19/Scotland/QEUH-B31821/2020\|EPI_ISL_676426\|2 | .................... | .................... | ..................... | .................... | .................... | .................... |
| U | 5444 hCoV-19/Scotland/QEUH-B3185E/2020\|EPI_ISL_676427\|2 | .................... | .................... | ..................... | .................... | .................... | .................... |
| U | 5445 hCoV-19/Scotland/QEUH-B31803/2020\|EPI_ISL_676428\|2 | .................... | .................... | ..................... | .................... | ......T............. | .................... |
| U | 5446 hCoV-19/Scotland/QEUH-B2B8EE/2020\|EPI_ISL_676429\|2 | .................... | .................... | ..................... | .................... | .................... | .................... |
| U | 5447 hCoV-19/Scotland/QEUH-B2B972/2020\|EPI_ISL_676430\|2 | .................... | .................... | ..................... | .................... | .................... | .................... |
| U | 5448 hCoV-19/Scotland/QEUH-B2B2A4/2020\|EPI_ISL_676432\|2 | .................... | .................... | ..................... | .................... | .................... | .................... |
| U | 5449 hCoV-19/Scotland/QEUH-B2B936/2020\|EPI_ISL_676434\|2 | .................... | .................... | ..................... | .................... | .................... | .................... |
| U | 5450 hCoV-19/Scotland/QEUH-B2B6C6/2020\|EPI_ISL_676435\|2 | .................... | .................... | ..................... | .................... | .................... | .................... |
| U | 5451 hCoV-19/Scotland/QEUH-B2B268/2020\|EPI_ISL_676436\|2 | .................... | .................... | ..................... | .................... | .................... | .................... |
| U | 5452 hCoV-19/Scotland/QEUH-B2B30B/2020\|EPI_ISL_676437\|2 | .................... | .................... | ..................... | .................... | .................... | .................... |
| U | 5453 hCoV-19/Scotland/QEUH-B2DB2F/2020\|EPI_ISL_676438\|2 | .................... | .................... | ..................... | .................... | ......T............. | .................... |
| U | 5454 hCoV-19/Scotland/QEUH-B2DCFF/2020\|EPI_ISL_676439\|2 | .................... | .................... | ..................... | .................... | .................... | .................... |
| U | 5455 hCoV-19/Scotland/QEUH-B2ED64/2020\|EPI_ISL_676440\|2 | .................... | .................... | ..................... | .................... | .................... | .................... |
| U | 5456 hCoV-19/Scotland/QEUH-AEFF00/2020\|EPI_ISL_676442\|2 | .................... | .................... | ..................... | .................... | .................... | .................... |
| U | 5457 hCoV-19/Scotland/QEUH-AEFF5B/2020\|EPI_ISL_676443\|2 | .................... | .................... | ..................... | .................... | .................... | .................... |
| U | 5458 hCoV-19/Scotland/QEUH-AF01D9/2020\|EPI_ISL_676444\|2 | .................... | .................... | ..................... | .................... | .................... | .................... |
| U | 5459 hCoV-19/Scotland/QEUH-AEFF6A/2020\|EPI_ISL_676446\|2 | .................... | .................... | ..................... | .................... | .................... | .................... |
| U | 5460 hCoV-19/Scotland/QEUH-AEFED6/2020\|EPI_ISL_676447\|2 | .................... | .................... | ..................... | .................... | .................... | .................... |
| U | 5461 hCoV-19/Scotland/QEUH-AEFEE5/2020\|EPI_ISL_676448\|2 | .................... | .................... | ..................... | .................... | .................... | .................... |
| U | 5462 hCoV-19/Scotland/QEUH-AEFF2E/2020\|EPI_ISL_676449\|2 | .................... | .A.................. | ..................... | .................... | .................... | .................... |
| U | 5463 hCoV-19/Scotland/QEUH-AEFEA9/2020\|EPI_ISL_676451\|2 | .................... | .................... | ..................... | .................... | .................... | .................... |
| U | 5464 hCoV-19/Scotland/QEUH-AEFF79/2020\|EPI_ISL_676452\|2 | .................... | .................... | ..................... | .................... | .................... | .................... |
| U | 5465 hCoV-19/Scotland/QEUH-AEFE6D/2020\|EPI_ISL_676453\|2 | .................... | .................... | ..................... | .................... | .................... | .................... |
| U | 5466 hCoV-19/Scotland/QEUH-AEFE5E/2020\|EPI_ISL_676455\|2 | .................... | .................... | ..................... | .................... | .................... | .................... |
| U | 5467 hCoV-19/Scotland/QEUH-AF2869/2020\|EPI_ISL_676456\|2 | .................... | .................... | ..................... | .................... | .................... | .................... |
| U | 5468 hCoV-19/Scotland/QEUH-AEFEB8/2020\|EPI_ISL_676457\|2 | .................... | .................... | ..................... | .................... | .................... | .................... |
| U | 5469 hCoV-19/Scotland/QEUH-AF47D3/2020\|EPI_ISL_676459\|2 | .................... | .................... | ..................... | .................... | .................... | .................... |
| U | 5470 hCoV-19/Scotland/QEUH-AF2887/2020\|EPI_ISL_676460\|2 | .................... | .................... | ..................... | .................... | .................... | .................... |
| U | 5471 hCoV-19/Scotland/QEUH-AEFF3D/2020\|EPI_ISL_676461\|2 | .................... | .................... | ..................... | .................... | .................... | .................... |
| U | 5472 hCoV-19/Scotland/QEUH-AF5356/2020\|EPI_ISL_676462\|2 | .................... | .................... | ..................... | .................... | .................... | .................... |
| U | 5473 hCoV-19/Scotland/QEUH-AF5DC1/2020\|EPI_ISL_676464\|2 | .................... | .................... | ..................... | .................... | .................... | .................... |
| U | 5474 hCoV-19/Scotland/QEUH-AF5AF7/2020\|EPI_ISL_676465\|2 | .................... | .................... | ..................... | .................... | .................... | .................... |
| U | 5475 hCoV-19/Scotland/QEUH-AF5277/2020\|EPI_ISL_676466\|2 | .................... | .................... | ..................... | .................... | .................... | .................... |
| U | 5476 hCoV-19/Scotland/QEUH-AF5BB8/2020\|EPI_ISL_676467\|2 | .................... | .................... | ..................... | .................... | .................... | .................... |
| U | 5477 hCoV-19/Scotland/QEUH-AF6258/2020\|EPI_ISL_676468\|2 | .................... | .................... | ..................... | .................... | .................... | .................... |
| U | 5478 hCoV-19/Scotland/QEUH-AF52D1/2020\|EPI_ISL_676469\|2 | .................... | .................... | ..................... | .................... | .................... | .................... |
| U | 5479 hCoV-19/Scotland/QEUH-AF516B/2020\|EPI_ISL_676470\|2 | .................... | .................... | ..................... | .................... | .................... | .................... |
| U | 5480 hCoV-19/Scotland/QEUH-AF5286/2020\|EPI_ISL_676471\|2 | .................... | .................... | ..................... | .................... | .................... | .................... |
| U | 5481 hCoV-19/Scotland/QEUH-AF50AA/2020\|EPI_ISL_676472\|2 | .................... | .................... | ..................... | .................... | .................... | .................... |
| U | 5482 hCoV-19/Scotland/QEUH-AF5110/2020\|EPI_ISL_676473\|2 | .................... | .................... | ..................... | .................... | .................... | .................... |
| U | 5483 hCoV-19/Scotland/QEUH-AF614C/2020\|EPI_ISL_676474\|2 | .................... | .................... | ..................... | .................... | .................... | .................... |
| U | 5484 hCoV-19/Scotland/QEUH-AF6188/2020\|EPI_ISL_676475\|2 | .................... | .................... | ..................... | .................... | .................... | .................... |
| U | 5485 hCoV-19/Scotland/QEUH-AF6689/2020\|EPI_ISL_676476\|2 | .................... | .................... | ..................... | .................... | .................... | .................... |
| U | 5486 hCoV-19/Scotland/QEUH-AF4B04/2020\|EPI_ISL_676477\|2 | .................... | .................... | ..................... | .................... | .................... | .................... |
| U | 5487 hCoV-19/Scotland/QEUH-AF51B6/2020\|EPI_ISL_676478\|2 | .................... | .................... | ..................... | .................... | .................... | .................... |
| U | 5488 hCoV-19/Scotland/QEUH-AF4D2C/2020\|EPI_ISL_676479\|2 | .................... | .................... | ..................... | .................... | .................... | .................... |
| U | 5489 hCoV-19/Scotland/QEUH-AF6179/2020\|EPI_ISL_676480\|2 | .................... | .................... | ..................... | .................... | .................... | .................... |
| U | 5490 hCoV-19/Scotland/QEUH-AF4E1A/2020\|EPI_ISL_676481\|2 | .................... | .................... | ..................... | .................... | .................... | .................... |
| U | 5491 hCoV-19/Scotland/QEUH-AF5DFE/2020\|EPI_ISL_676482\|2 | .................... | .................... | ..................... | .................... | .................... | .................... |
| U | 5492 hCoV-19/Scotland/QEUH-AF6197/2020\|EPI_ISL_676483\|2 | .................... | .................... | ..................... | .................... | .................... | .................... |
| U | 5493 hCoV-19/Scotland/QEUH-AF4D95/2020\|EPI_ISL_676485\|2 | .................... | .................... | ..................... | .................... | .................... | .................... |
| U | 5494 hCoV-19/Scotland/QEUH-AF4ADA/2020\|EPI_ISL_676486\|2 | .................... | .................... | ..................... | .................... | .................... | .................... |
| U | 5495 hCoV-19/Scotland/QEUH-AF4B8C/2020\|EPI_ISL_676487\|2 | .................... | .................... | ..................... | .................... | .................... | .................... |
| U | 5496 hCoV-19/Scotland/QEUH-AF61E2/2020\|EPI_ISL_676488\|2 | .................... | .................... | ..................... | .................... | .................... | .................... |
| U | 5497 hCoV-19/Sweden/20-09868/2020\|EPI_ISL_676491\|2020-1 | .................... | .................... | ..................... | .................... | .................... | .................... |
| U | 5498 hCoV-19/Sweden/20-52780/2020\|EPI_ISL_676492\|2020-0 | .................... | .................... | ..................... | .................... | .................... | .................... |
| U | 5499 hCoV-19/Sweden/20-53008/2020\|EPI_ISL_676493\|2020-0 | .................... | .................... | ..................... | .................... | .................... | .................... |
| U | 5500 hCoV-19/Sweden/20-53077/2020\|EPI_ISL_676494\|2020-1 | .................... | .................... | ..................... | .................... | .................... | .................... |
| U | 5501 hCoV-19/Sweden/20-53093/2020\|EPI_ISL_676495\|2020-0 | .................... | .................... | ..................... | .................... | .................... | .................... |
| U | 5502 hCoV-19/Sweden/20-53236/2020\|EPI_ISL_676496\|2020-1 | .................... | .................... | ..................... | .................... | .................... | .................... |
| U | 5503 hCoV-19/Sweden/20-53395/2020\|EPI_ISL_676497\|2020-1 | .................... | .................... | ..................... | .................... | .................... | .................... |
| U | 5504 hCoV-19/Sweden/20-53396/2020\|EPI_ISL_676498\|2020-1 | .................... | .................... | ..................... | .................... | .................... | .................... |
| U | 5505 hCoV-19/Sweden/20-53397/2020\|EPI_ISL_676499\|2020-1 | .................... | .................... | ..................... | .................... | .................... | .................... |
| U | 5506 hCoV-19/Sweden/20-53399/2020\|EPI_ISL_676500\|2020-1 | .................... | .................... | ..................... | .................... | .................... | .................... |
| U | 5507 hCoV-19/Sweden/20-53400/2020\|EPI_ISL_676501\|2020-1 | .................... | .................... | ..................... | .................... | .................... | .................... |

| | | | | | | | | |
|---|---|---|---|---|---|---|---|---|
| U | 5508 hCoV-19/Hungary/US-32521w/2020\|EPI_ISL_677750\|2020 | .................. | .................. | .................. | .................. | .................. | .................. | |
| U | 5509 hCoV-19/Hungary/US-32533w,/2020\|EPI_ISL_677751\|202 | .................. | .................. | .................. | .................. | .................. | .................. | |
| U | 5510 hCoV-19/Hungary/US-32534w/2020\|EPI_ISL_677752\|2020 | .................. | .................. | .................. | .................. | .................. | .................. | |
| U | 5511 hCoV-19/Hungary/US-32539w/2020\|EPI_ISL_677753\|2020 | .................. | .................. | .................. | .................. | .................. | .................. | |
| U | 5512 hCoV-19/Hungary/US-32557w/2020\|EPI_ISL_677756\|2020 | .................. | .................. | .................. | .................. | .................. | .................. | |
| U | 5513 hCoV-19/Hungary/US-32566w/2020\|EPI_ISL_677757\|2020 | .................. | .................. | .................. | .................. | .................. | .................. | |
| U | 5514 hCoV-19/Hungary/US-32570w/2020\|EPI_ISL_677758\|2020 | .................. | .................. | .................. | .................. | .................. | .................. | |
| U | 5515 hCoV-19/Hungary/US-32571w/2020\|EPI_ISL_677759\|2020 | .................. | .................. | .................. | .................. | .................. | .................. | |
| U | 5516 hCoV-19/Hungary/US-32620w/2020\|EPI_ISL_677761\|2020 | .................. | .................. | .................. | .................. | .................. | .................. | |
| U | 5517 hCoV-19/Hungary/US-32637w/2020\|EPI_ISL_677762\|2020 | .................. | .................. | .................. | .................. | .................. | .................. | |
| U | 5518 hCoV-19/Hungary/US-33714w/2020\|EPI_ISL_677763\|2020 | .................. | .................. | .................. | .................. | .................. | .................. | |
| U | 5519 hCoV-19/Hungary/US-33735w/2020\|EPI_ISL_677764\|2020 | .................. | .................. | .................. | .................. | .................. | .................. | |
| U | 5520 hCoV-19/Hungary/US-33736w/2020\|EPI_ISL_677765\|2020 | .................. | .................. | .................. | .................. | .................. | .................. | |
| U | 5521 hCoV-19/Hungary/US-33755w/2020\|EPI_ISL_677766\|2020 | .................. | .................. | .................. | .................. | .................. | .................. | |
| U | 5522 hCoV-19/Hungary/US-33782w/2020\|EPI_ISL_677767\|2020 | .................. | .................. | .................. | .................. | .................. | .................. | |
| U | 5523 hCoV-19/Hungary/US-35090w/2020\|EPI_ISL_677768\|2020 | .................. | .................. | .................. | .................. | .................. | .................. | |
| U | 5526 hCoV-19/Hungary/US-55491w/2020\|EPI_ISL_677773\|2020 | .................. | .................. | .................. | .................. | .................. | .................. | |
| U | 5527 hCoV-19/Hungary/US-55497w/2020\|EPI_ISL_677774\|2020 | .................. | .................. | .................. | .................. | .................. | .................. | |
| U | 5528 hCoV-19/Hungary/US-56260w/2020\|EPI_ISL_677775\|2020 | .................. | .................. | .................. | .................. | .................. | .................. | |
| U | 5529 hCoV-19/Hungary/US-56267w/2020\|EPI_ISL_677776\|2020 | .................. | .................. | .................. | .................. | .................. | .................. | |
| U | 5530 hCoV-19/Hungary/US-57376w/2020\|EPI_ISL_677777\|2020 | .................. | .................. | .................. | .................. | .................. | .................. | |
| U | 5531 hCoV-19/Hungary/US-57417w/2020\|EPI_ISL_677780\|2020 | .................. | .................. | .................. | .................. | .................. | .................. | |
| U | 5533 hCoV-19/Hungary/US-57479w/2020\|EPI_ISL_677782\|2020 | .................. | .................. | .................. | .................. | .................. | .................. | |
| U | 5534 hCoV-19/Hungary/US-57508w/2020\|EPI_ISL_677783\|2020 | .................. | .................. | .................. | .................. | .................. | .................. | |
| U | 5536 hCoV-19/Hungary/US-57524w/2020\|EPI_ISL_677786\|2020 | .................. | .................. | .................. | .................. | .................. | .................. | |
| U | 5537 hCoV-19/Hungary/US-57525w/2020\|EPI_ISL_677787\|2020 | .................. | .................. | .................. | .................. | .................. | .................. | |
| U | 5538 hCoV-19/Hungary/US-57527w/2020\|EPI_ISL_677788\|2020 | .................. | .................. | .................. | .................. | .................. | .................. | |
| U | 5539 hCoV-19/Hungary/US-57529w/2020\|EPI_ISL_677789\|2020 | .................. | .................. | .................. | .................. | .................. | .................. | |
| U | 5540 hCoV-19/Hungary/US-57540w/2020\|EPI_ISL_677790\|2020 | .................. | .................. | .................. | .................. | .................. | .................. | |
| U | 5541 hCoV-19/Hungary/US-57543w/2020\|EPI_ISL_677791\|2020 | .................. | .................. | .................. | .................. | .................. | .................. | |
| U | 5542 hCoV-19/Hungary/US-57552w/2020\|EPI_ISL_677792\|2020 | .................. | .................. | .................. | .................. | .................. | .................. | |
| U | 5543 hCoV-19/Hungary/US-57557w/2020\|EPI_ISL_677793\|2020 | .................. | .................. | .................. | .................. | .................. | .................. | |
| U | 5544 hCoV-19/Hungary/US-57565w/2020\|EPI_ISL_677794\|2020 | .................. | .................. | .................. | .................. | .................. | .................. | |
| U | 5545 hCoV-19/Hungary/US-57566w/2020\|EPI_ISL_677795\|2020 | .................. | .................. | .................. | .................. | .................. | .................. | |
| U | 5546 hCoV-19/Hungary/US-57572w/2020\|EPI_ISL_677796\|2020 | .................. | .................. | .................. | .................. | .................. | .................. | |
| U | 5547 hCoV-19/Hungary/US-57574w/2020\|EPI_ISL_677797\|2020 | .................. | .................. | .................. | .................. | .................. | .................. | |
| U | 5548 hCoV-19/Hungary/US-57575w/2020\|EPI_ISL_677798\|2020 | .................. | .................. | .................. | .................. | .................. | .................. | |
| U | 5549 hCoV-19/Hungary/US-57576w/2020\|EPI_ISL_677799\|2020 | .................. | .................. | .................. | .................. | .................. | .................. | |
| U | 5550 hCoV-19/Hungary/US-57595w/2020\|EPI_ISL_677800\|2020 | .................. | .................. | .................. | .................. | .................. | .................. | |
| U | 5551 hCoV-19/Hungary/US-60346w/2020\|EPI_ISL_677801\|2020 | .................. | .................. | .................. | .................. | .................. | .................. | |
| U | 5552 hCoV-19/Hungary/US-60347w/2020\|EPI_ISL_677802\|2020 | .................. | .................. | .................. | .................. | .................. | .................. | |
| U | 5553 hCoV-19/Hungary/US-60377w/2020\|EPI_ISL_677805\|2020 | .................. | .................. | .................. | .................. | .................. | .................. | |
| U | 5554 hCoV-19/Hungary/US-60431w/2020\|EPI_ISL_677806\|2020 | .................. | .................. | .................. | .................. | .................. | .................. | |
| U | 5555 hCoV-19/Hungary/US-60434w/2020\|EPI_ISL_677808\|2020 | .................. | .................. | .................. | .................. | .................. | .................. | |
| U | 5556 hCoV-19/Hungary/US-60435w/2020\|EPI_ISL_677809\|2020 | .................. | .................. | .................. | .................. | .................. | .................. | |
| U | 5557 hCoV-19/Hungary/US-60505w/2020\|EPI_ISL_677810\|2020 | .................. | .................. | .................. | .................. | .................. | .................. | |
| U | 5558 hCoV-19/Hungary/US-60522w/2020\|EPI_ISL_677811\|2020 | .................. | .................. | .................. | .................. | .................. | .................. | |
| U | 5559 hCoV-19/Hungary/US-60527w/2020\|EPI_ISL_677812\|2020 | .................. | .................. | .................. | .................. | ................C. | .................. | 1 |
| U | 5560 hCoV-19/Hungary/US-60541w/2020\|EPI_ISL_677813\|2020 | .................. | .................. | .................. | .................. | .................. | .................. | |
| U | 5561 hCoV-19/Hungary/US-60544w/2020\|EPI_ISL_677814\|2020 | .................. | .................. | .................. | .................. | .................. | .................. | |
| U | 5562 hCoV-19/Hungary/US-60568w/2020\|EPI_ISL_677815\|2020 | .................. | .................. | .................. | .................. | .................. | .................. | |
| U | 5563 hCoV-19/Hungary/US-60592w/2020\|EPI_ISL_677816\|2020 | .................. | .................. | .................. | .................. | .................. | .................. | |
| U | 5564 hCoV-19/Hungary/US-60595w/2020\|EPI_ISL_677817\|2020 | .................. | .................. | .................. | .................. | .................. | .................. | |
| U | 5565 hCoV-19/Hungary/US-63527w/2020\|EPI_ISL_677818\|2020 | .................. | .................. | .................. | .................. | .................. | .................. | |
| U | 5567 hCoV-19/USA/CA-IGI-0173/2020\|EPI_ISL_677821\|2020-0 | .................. | .................. | .................. | .................. | .................. | .................. | |
| U | 5568 hCoV-19/USA/CA-IGI-0424/2020\|EPI_ISL_677824\|2020-0 | .................. | .A................ | .................. | .................. | .................. | .................. | |
| U | 5569 hCoV-19/USA/CA-IGI-0442/2020\|EPI_ISL_677825\|2020-0 | .................. | .................. | .................. | .................. | .................. | .................. | |
| U | 5570 hCoV-19/USA/CA-IGI-0546/2020\|EPI_ISL_677827\|2020-0 | .................. | .................. | .................. | .................. | .................. | .................. | |
| U | 5571 hCoV-19/USA/CA-IGI-0564/2020\|EPI_ISL_677832\|2020-0 | .................. | .................. | .................. | .................. | .................. | .................. | |
| U | 5572 hCoV-19/USA/CA-IGI-0593/2020\|EPI_ISL_677839\|2020-0 | .................. | .................. | .................. | .................. | .................. | .................. | |
| U | 5573 hCoV-19/Wales/PHWC-48F0F6/2020\|EPI_ISL_681045\|2020 | .................. | .................. | .................. | .................. | .................. | .................. | |
| U | 5574 hCoV-19/Wales/PHWC-48F111/2020\|EPI_ISL_681047\|2020 | .................. | .................. | .................. | .................. | .................. | .................. | |
| U | 5577 hCoV-19/Wales/PHWC-48F16C/2020\|EPI_ISL_681051\|2020 | .................. | .................. | .................. | .................. | .................. | .................. | |
| U | 5578 hCoV-19/Wales/PHWC-48F18A/2020\|EPI_ISL_681052\|2020 | .................. | .................. | .................. | .................. | .................. | .................. | |
| U | 5580 hCoV-19/Wales/PHWC-48F1A8/2020\|EPI_ISL_681054\|2020 | .................. | .................. | .................. | .................. | .................. | .................. | |
| U | 5581 hCoV-19/Wales/PHWC-48F1B7/2020\|EPI_ISL_681055\|2020 | .................. | .................. | .................. | .................. | .................. | .................. | |
| U | 5582 hCoV-19/Wales/PHWC-48F1C6/2020\|EPI_ISL_681056\|2020 | .................. | .................. | .................. | .................. | .................. | .................. | |
| U | 5583 hCoV-19/Wales/PHWC-48F1D5/2020\|EPI_ISL_681057\|2020 | .................. | .................. | .................. | .................. | .................. | .................. | |
| U | 5584 hCoV-19/Wales/PHWC-48F1E4/2020\|EPI_ISL_681058\|2020 | .................. | .................. | .................. | .................. | .................. | .................. | |
| U | 5585 hCoV-19/Wales/PHWC-48F1F3/2020\|EPI_ISL_681059\|2020 | .................. | .................. | .................. | .................. | .................. | .................. | |
| U | 5586 hCoV-19/Wales/PHWC-48F20F/2020\|EPI_ISL_681060\|2020 | .................. | .................. | .................. | .................. | .................. | .................. | |
| U | 5587 hCoV-19/Wales/PHWC-48F21E/2020\|EPI_ISL_681061\|2020 | .................. | .................. | .................. | .................. | .................. | .................. | |
| U | 5588 hCoV-19/Wales/PHWC-48F269/2020\|EPI_ISL_681065\|2020 | .................. | .................. | .................. | .................. | .................. | .................. | |
| U | 5589 hCoV-19/Wales/PHWC-48F278/2020\|EPI_ISL_681066\|2020 | .................. | .................. | .................. | .................. | .................. | .................. | |
| U | 5590 hCoV-19/Wales/PHWC-48F2A5/2020\|EPI_ISL_681069\|2020 | .................. | .................. | .................. | .................. | .................. | .................. | |
| U | 5591 hCoV-19/Wales/PHWC-48F2B4/2020\|EPI_ISL_681070\|2020 | .................. | .................. | .................. | .................. | .................. | .................. | |
| U | 5592 hCoV-19/Wales/PHWC-48F2C3/2020\|EPI_ISL_681071\|2020 | .................. | .................. | .................. | .................. | .................. | .................. | |
| U | 5593 hCoV-19/Wales/PHWC-48F2D2/2020\|EPI_ISL_681072\|2020 | .................. | .................. | .................. | .................. | .................. | .................. | |
| U | 5594 hCoV-19/Wales/PHWC-48F2E1/2020\|EPI_ISL_681073\|2020 | .................. | .................. | .................. | .................. | .................. | .................. | |
| U | 5595 hCoV-19/Wales/PHWC-48F2F0/2020\|EPI_ISL_681074\|2020 | .................. | .................. | .................. | .................. | .................. | .................. | |

U 5597 hCoV-19/Wales/PHWC-48F31B/2020|EPI_ISL_681076|2020 ···················· ···················· ····················· ···················· ···················· ····················
U 5598 hCoV-19/Wales/PHWC-48F348/2020|EPI_ISL_681078|2020 ···················· ···················· ····················· ···················· ···················· ····················
U 5599 hCoV-19/Wales/PHWC-48F357/2020|EPI_ISL_681079|2020 ···················· ···················· ····················· ···················· ···················· ····················
U 5600 hCoV-19/Wales/PHWC-48F366/2020|EPI_ISL_681080|2020 ···················· ······R············· ····················· ···················· ···················· ····················
U 5602 hCoV-19/Wales/PHWC-48F384/2020|EPI_ISL_681082|2020 ···················· ···················· ····················· ···················· ···················· ····················
U 5603 hCoV-19/Wales/PHWC-48F393/2020|EPI_ISL_681083|2020 ···················· ···················· ····················· ···················· ···················· ····················
U 5604 hCoV-19/Wales/PHWC-48F3A2/2020|EPI_ISL_681084|2020 ···················· ···················· ····················· ···················· ···················· ····················
U 5605 hCoV-19/Wales/PHWC-48F3B1/2020|EPI_ISL_681085|2020 ···················· ···················· ····················· ···················· ···················· ····················
U 5606 hCoV-19/Wales/PHWC-48F3C0/2020|EPI_ISL_681086|2020 ···················· ······A············· ····················· ···················· ···················· ····················
U 5607 hCoV-19/Wales/PHWC-48F3DF/2020|EPI_ISL_681087|2020 ···················· ······A············· ····················· ···················· ···················· ····················
U 5608 hCoV-19/Wales/PHWC-48F3EE/2020|EPI_ISL_681088|2020 ···················· ···················· ····················· ···················· ···················· ····················
U 5609 hCoV-19/Wales/PHWC-48F409/2020|EPI_ISL_681090|2020 ···················· ···················· ····················· ···················· ···················· ····················
U 5610 hCoV-19/Wales/PHWC-48F418/2020|EPI_ISL_681091|2020 ···················· ···················· ····················· ···················· ···················· ····················
U 5611 hCoV-19/Wales/PHWC-48F445/2020|EPI_ISL_681093|2020 ···················· ···················· ····················· ···················· ···················· ····················
U 5612 hCoV-19/Wales/PHWC-48F454/2020|EPI_ISL_681094|2020 ···················· ···················· ····················· ···················· ···················· ····················
U 5613 hCoV-19/Wales/PHWC-48F463/2020|EPI_ISL_681095|2020 ···················· ···················· ····················· ···················· ···················· ····················
U 5615 hCoV-19/Wales/PHWC-48F481/2020|EPI_ISL_681097|2020 ···················· ···················· ····················· ···················· ···················· ····················
U 5617 hCoV-19/Wales/PHWC-48F4AF/2020|EPI_ISL_681099|2020 ···················· ···················· ····················· ···················· ···················· ····················
U 5618 hCoV-19/Denmark/DCGC-12197/2020|EPI_ISL_682378|202 ···················· ···················· ····················· ···················· ···················· ····················
U 5619 hCoV-19/Denmark/DCGC-12198/2020|EPI_ISL_682379|202 ···················· ···················· ····················· ···················· ···················· ····················
U 5620 hCoV-19/Denmark/DCGC-12199/2020|EPI_ISL_682380|202 ···················· ···················· ····················· ···················· ···················· ····················
U 5621 hCoV-19/Denmark/DCGC-12200/2020|EPI_ISL_682381|202 ···················· ···················· ····················· ···················· ···················· ····················
U 5622 hCoV-19/Denmark/DCGC-12201/2020|EPI_ISL_682382|202 ···················· ···················· ····················· ···················· ···················· ····················
U 5623 hCoV-19/Denmark/DCGC-12202/2020|EPI_ISL_682383|202 ···················· ···················· ····················· ···················· ···················· ····················
U 5624 hCoV-19/Denmark/DCGC-12203/2020|EPI_ISL_682384|202 ···················· ···················· ····················· ···················· ······T············· ····················
U 5625 hCoV-19/Denmark/DCGC-12204/2020|EPI_ISL_682385|202 ···················· ···················· ····················· ···················· ···················· ····················
U 5626 hCoV-19/Denmark/DCGC-12205/2020|EPI_ISL_682386|202 ···················· ·A·················· ····················· ···················· ···················· ···A················
U 5627 hCoV-19/Denmark/DCGC-12206/2020|EPI_ISL_682387|202 ···················· ···················· ····················· ···················· ···················· ····················
U 5628 hCoV-19/Denmark/DCGC-12208/2020|EPI_ISL_682389|202 ···················· ···················· ····················· ···················· ···················· ····················
U 5629 hCoV-19/Denmark/DCGC-12210/2020|EPI_ISL_682391|202 ···················· ·A·················· ····················· ···················· ···················· ····················
U 5630 hCoV-19/Denmark/DCGC-12214/2020|EPI_ISL_682395|202 ···················· ···················· ····················· ···················· ···················· ····················
U 5631 hCoV-19/Denmark/DCGC-12216/2020|EPI_ISL_682397|202 ···················· ···················· ····················· ···················· ···················· ····················
U 5632 hCoV-19/Denmark/DCGC-12217/2020|EPI_ISL_682398|202 ···················· ···················· ····················· ···················· ···················· ····················
U 5633 hCoV-19/England/QEUH-BA265E/2020|EPI_ISL_672900|20 ···················· ···················· ····················· ···················· ···················· ····················
U 5634 hCoV-19/England/QEUH-BA2ABC/2020|EPI_ISL_672902|20 ···················· ···················· ····················· ···················· ···················· ····················
U 5635 hCoV-19/England/QEUH-BA2A16/2020|EPI_ISL_672903|20 ···················· ···················· ····················· ···················· ···················· ····················
U 5636 hCoV-19/England/QEUH-BA3435/2020|EPI_ISL_672904|20 ···················· ···················· ····················· ···················· ···················· ····················
U 5637 hCoV-19/England/QEUH-BA1200/2020|EPI_ISL_672905|20 ···················· ···················· ····················· ···················· ···················· ····················
U 5638 hCoV-19/England/QEUH-BA2E65/2020|EPI_ISL_672907|20 ···················· ···················· ····················· ···················· ···················· ····················
U 5639 hCoV-19/England/QEUH-BA1F09/2020|EPI_ISL_672908|20 ···················· ···················· ····················· ···················· ···················· ····················
U 5640 hCoV-19/England/QEUH-BA313E/2020|EPI_ISL_672911|20 ···················· ···················· ····················· ···················· ···················· ····················
U 5641 hCoV-19/England/CAMC-B0993E/2020|EPI_ISL_672912|20 ···················· ···················· ····················· ···················· ···················· ····················
U 5642 hCoV-19/England/CAMC-B09A86/2020|EPI_ISL_672913|20 ···················· ···················· ····················· ···················· ······T············· ····················
U 5643 hCoV-19/England/CAMC-B09A95/2020|EPI_ISL_672914|20 ···················· ···················· ····················· ···················· ···················· ····················
U 5644 hCoV-19/England/CAMC-B09998/2020|EPI_ISL_672915|20 ···················· ··············C····· ····················· ···················· ···················· ····················
U 5645 hCoV-19/England/CAMC-B09E3F/2020|EPI_ISL_672916|20 ···················· ···················· ····················· ···················· ···················· ····················
U 5646 hCoV-19/England/CAMC-B09619/2020|EPI_ISL_672917|20 ···················· ···················· ····················· ···················· ···················· ····················
U 5647 hCoV-19/England/CAMC-B099D4/2020|EPI_ISL_672918|20 ···················· ···················· ····················· ···················· ···················· ····················
U 5648 hCoV-19/England/CAMC-B09CDB/2020|EPI_ISL_672919|20 ···················· ···················· ····················· ···················· ···················· ····················
U 5649 hCoV-19/England/CAMC-B09AFF/2020|EPI_ISL_672920|20 ···················· ···················· ····················· ···················· ···················· ····················
U 5650 hCoV-19/England/CAMC-B0A038/2020|EPI_ISL_672921|20 ···················· ···················· ····················· ···················· ···················· ····················
U 5651 hCoV-19/England/CAMC-B09057/2020|EPI_ISL_672922|20 ···················· ···················· ····················· ···················· ···················· ····················
U 5652 hCoV-19/England/CAMC-B08E12/2020|EPI_ISL_672923|20 ···················· ···················· ····················· ···················· ···················· ····················
U 5653 hCoV-19/England/CAMC-B08E6D/2020|EPI_ISL_672925|20 ···················· ···················· ····················· ···················· ···················· ····················
U 5654 hCoV-19/England/CAMC-B09C08/2020|EPI_ISL_672926|20 ···················· ···················· ····················· ···················· ···················· ····················
U 5655 hCoV-19/England/CAMC-B09B74/2020|EPI_ISL_672927|20 ···················· ···················· ····················· ···················· ···················· ····················
U 5656 hCoV-19/England/CAMC-B0A223/2020|EPI_ISL_672928|20 ···················· ···················· ····················· ···················· ···················· ····················
U 5657 hCoV-19/England/CAMC-B09FD2/2020|EPI_ISL_672929|20 ···················· ···················· ····················· ···················· ······T············· ····················
U 5658 hCoV-19/England/CAMC-B09AC2/2020|EPI_ISL_672930|20 ···················· ···················· ····················· ···················· ···················· ····················
U 5659 hCoV-19/England/CAMC-B095B2/2020|EPI_ISL_672931|20 ···················· ·A·················· ···········T········· ···················· ···················· ···A················
U 5660 hCoV-19/England/CAMC-B09C53/2020|EPI_ISL_672932|20 ···················· ···················· ····················· ···················· ······T············· ····················
U 5661 hCoV-19/England/CAMC-B09BFC/2020|EPI_ISL_672933|20 ···················· ···················· ····················· ···················· ···················· ····················
U 5662 hCoV-19/England/CAMC-B0A083/2020|EPI_ISL_672934|20 ···················· ···················· ····················· ···················· ···················· ····················
U 5663 hCoV-19/England/CAMC-B09A68/2020|EPI_ISL_672935|20 ···················· ···················· ····················· ···················· ···················· ····················
U 5664 hCoV-19/England/CAMC-B0A029/2020|EPI_ISL_672936|20 ···················· ···················· ····················· ···················· ······T············· ····················
U 5665 hCoV-19/England/CAMC-B0979E/2020|EPI_ISL_672937|20 ···················· ···················· ····················· ···················· ···················· ····················
U 5666 hCoV-19/England/CAMC-B096DD/2020|EPI_ISL_672938|20 ···················· ···················· ····················· ···················· ···················· ····················
U 5667 hCoV-19/England/CAMC-B08BC0/2020|EPI_ISL_672941|20 ···················· ···················· ····················· ···················· ···················· ····················
U 5668 hCoV-19/England/CAMC-B09901/2020|EPI_ISL_672942|20 ···················· ···················· ····················· ···················· ···················· ····················
U 5669 hCoV-19/England/CAMC-B09330/2020|EPI_ISL_672943|20 ···················· ···················· ····················· ···················· ···················· ····················
U 5670 hCoV-19/England/CAMC-B090FD/2020|EPI_ISL_672945|20 ···················· ···················· ····················· ···················· ···················· ····················
U 5671 hCoV-19/England/CAMC-B09752/2020|EPI_ISL_672946|20 ···················· ···················· ····················· ···················· ···················· ····················
U 5672 hCoV-19/England/CAMC-B0902A/2020|EPI_ISL_672947|20 ···················· ···················· ····················· ···················· ···················· ····················
U 5673 hCoV-19/England/CAMC-B09BA1/2020|EPI_ISL_672948|20 ···················· ···················· ····················· ···················· ···················· ····················
U 5674 hCoV-19/England/CAMC-B09163/2020|EPI_ISL_672950|20 ···················· ···················· ····················· ···················· ······T············· ····················
U 5675 hCoV-19/England/CAMC-B095A3/2020|EPI_ISL_672952|20 ···················· ·A·················· ···········T········· ···················· ···················· ···A················
U 5676 hCoV-19/England/CAMC-B09DE7/2020|EPI_ISL_672953|20 ···················· ···················· ····················· ···················· ···················· ····················
U 5677 hCoV-19/England/CAMC-B09F1E/2020|EPI_ISL_672955|20 ···················· ···················· ····················· ···················· ···················· ····················
U 5678 hCoV-19/England/CAMC-B08FC4/2020|EPI_ISL_672956|20 ···················· ···················· ····················· ···················· ···················· ····················
U 5679 hCoV-19/England/CAMC-B097F8/2020|EPI_ISL_672957|20 ···················· ···················· ····················· ···················· ···················· ····················

| | | | | | | | |
|---|---|---|---|---|---|---|---|
| U | 5680 hCoV-19/England/CAMC-B09E02/2020\|EPI_ISL_672958\|20 | ···················· | ···················· | ···················· | ···················· | ···················· | ···················· |
| U | 5681 hCoV-19/England/CAMC-B08EA9/2020\|EPI_ISL_672959\|20 | ···················· | ···················· | ···················· | ···················· | ···················· | ···················· |
| U | 5682 hCoV-19/England/CAMC-B09637/2020\|EPI_ISL_672960\|20 | ···················· | ···················· | ···················· | ···················· | ···················· | ···················· |
| U | 5683 hCoV-19/England/CAMC-B09E6C/2020\|EPI_ISL_672961\|20 | ···················· | ···················· | ···················· | ···················· | ···················· | ···················· |
| U | 5684 hCoV-19/England/CAMC-B0A618/2020\|EPI_ISL_672963\|20 | ···················· | ···················· | ···················· | ···················· | ···················· | ···················· |
| U | 5685 hCoV-19/England/CAMC-B090B1/2020\|EPI_ISL_672964\|20 | ···················· | ···················· | ···················· | ···················· | ···················· | ···················· |
| U | 5686 hCoV-19/England/CAMC-B091AF/2020\|EPI_ISL_672965\|20 | ···················· | ···················· | ···················· | ···················· | ···················· | ···················· |
| U | 5687 hCoV-19/England/CAMC-B08F00/2020\|EPI_ISL_672966\|20 | ···················· | ···················· | ···················· | ···················· | ···················· | ···················· |
| U | 5688 hCoV-19/England/CAMC-B096BF/2020\|EPI_ISL_672967\|20 | ···················· | ···················· | ···················· | ···················· | ···················· | ···················· |
| U | 5689 hCoV-19/England/CAMC-B091FA/2020\|EPI_ISL_672968\|20 | ···················· | ···················· | ···················· | ···················· | ···················· | ···················· |
| U | 5690 hCoV-19/England/CAMC-B0A117/2020\|EPI_ISL_672969\|20 | ···················· | ···················· | ···················· | ···················· | ···················· | ···················· |
| U | 5691 hCoV-19/England/CAMC-B08EF4/2020\|EPI_ISL_672970\|20 | ···················· | ···················· | ···················· | ···················· | ···················· | ···················· |
| U | 5692 hCoV-19/England/CAMC-B99D46/2020\|EPI_ISL_673300\|20 | ···················· | ···················· | ···················· | ···················· | ···················· | ···················· |
| U | 5693 hCoV-19/England/CAMC-B997C0/2020\|EPI_ISL_673356\|20 | ···················· | ···················· | ···················· | ···················· | ···················· | ···················· |
| U | 5694 hCoV-19/England/CAMC-B99423/2020\|EPI_ISL_673357\|20 | ···················· | ···················· | ···················· | ···················· | ···················· | ···················· |
| U | 5695 hCoV-19/England/CAMC-B9AC48/2020\|EPI_ISL_673358\|20 | ···················· | ···················· | ···················· | ···················· | ···················· | ···················· |
| U | 5696 hCoV-19/England/CAMC-B99C1C/2020\|EPI_ISL_673365\|20 | ···················· | ···················· | ···················· | ···················· | ···················· | ···················· |
| U | 5697 hCoV-19/England/CAMC-B999F7/2020\|EPI_ISL_673368\|20 | ···················· | ···················· | ···················· | ···················· | ···················· | ···················· |
| U | 5698 hCoV-19/England/CAMC-B98FBA/2020\|EPI_ISL_673372\|20 | ···················· | ···················· | ···················· | ···················· | ···················· | ···················· |
| U | 5699 hCoV-19/England/CAMC-B991B3/2020\|EPI_ISL_673373\|20 | ···················· | ···················· | ···················· | ···················· | ···················· | ···················· |
| U | 5700 hCoV-19/England/CAMC-B9ABC3/2020\|EPI_ISL_673374\|20 | ···················· | ···················· | ···················· | ···················· | ···················· | ···················· |
| U | 5701 hCoV-19/England/CAMC-B98FD8/2020\|EPI_ISL_673375\|20 | ···················· | ···················· | ···················· | ···················· | ···················· | ···················· |
| U | 5702 hCoV-19/England/CAMC-B9AFF4/2020\|EPI_ISL_673376\|20 | ···················· | ···················· | ···················· | ···················· | ·····T·············· | ···················· |
| U | 5703 hCoV-19/England/CAMC-B99001/2020\|EPI_ISL_673377\|20 | ···················· | ···················· | ···················· | ···················· | ···················· | ···················· |
| U | 5704 hCoV-19/England/CAMC-B992A1/2020\|EPI_ISL_673378\|20 | ···················· | ···················· | ···················· | ···················· | ···················· | ···················· |
| U | 5705 hCoV-19/England/CAMC-B99C76/2020\|EPI_ISL_673379\|20 | ···················· | ···················· | ···················· | ···················· | ···················· | ···················· |
| U | 5706 hCoV-19/England/CAMC-B9954E/2020\|EPI_ISL_673380\|20 | ···················· | ···················· | ···················· | ···················· | ···················· | ···················· |
| U | 5707 hCoV-19/England/CAMC-B9A158/2020\|EPI_ISL_673381\|20 | ···················· | ···················· | ···················· | ···················· | ···················· | ··A················· |
| U | 5708 hCoV-19/England/CAMC-B99D37/2020\|EPI_ISL_673383\|20 | ···················· | ···················· | ···················· | ···················· | ···················· | ···················· |
| U | 5709 hCoV-19/England/CAMC-B9AC57/2020\|EPI_ISL_673384\|20 | ···················· | ···················· | ···················· | ···················· | ···················· | ···················· |
| U | 5710 hCoV-19/England/CAMC-B9A264/2020\|EPI_ISL_673385\|20 | ···················· | ···················· | ···················· | ···················· | ···················· | ···················· |
| U | 5711 hCoV-19/England/CAMC-B990A7/2020\|EPI_ISL_673386\|20 | ···················· | ···················· | ···················· | ···················· | ···················· | ···················· |
| U | 5712 hCoV-19/England/CAMC-B99CA3/2020\|EPI_ISL_673387\|20 | ···················· | ···················· | ···················· | ···················· | ···················· | ···················· |
| U | 5713 hCoV-19/England/CAMC-B99757/2020\|EPI_ISL_673388\|20 | ···················· | ···················· | ···················· | ···················· | ···················· | ···················· |
| U | 5714 hCoV-19/England/MILK-B8E0D2/2020\|EPI_ISL_674600\|20 | ···················· | ···················· | ···················· | ···················· | ···················· | ···················· |
| U | 5715 hCoV-19/England/MILK-B8DFF4/2020\|EPI_ISL_674601\|20 | ···················· | ···················· | ···················· | ···················· | ···················· | ···················· |
| U | 5716 hCoV-19/England/MILK-B8E00F/2020\|EPI_ISL_674602\|20 | ···················· | ···················· | ···················· | ···················· | ···················· | ···················· |
| U | 5717 hCoV-19/England/QEUH-B7F881/2020\|EPI_ISL_674603\|20 | ···················· | ···················· | ···················· | ···················· | ···················· | ···················· |
| U | 5718 hCoV-19/England/MILK-B8F4F3/2020\|EPI_ISL_674604\|20 | ···················· | ···················· | ···················· | ···················· | ·····T·············· | ···················· |
| U | 5719 hCoV-19/England/QEUH-B7F784/2020\|EPI_ISL_674605\|20 | ···················· | ···················· | ···················· | ···················· | ···················· | ···················· |
| U | 5720 hCoV-19/England/MILK-B8DF03/2020\|EPI_ISL_674606\|20 | ···················· | ···················· | ···················· | ···················· | ···················· | ···················· |
| U | 5721 hCoV-19/England/MILK-B8E04B/2020\|EPI_ISL_674608\|20 | ···················· | ···················· | ···················· | ···················· | ···················· | ···················· |
| U | 5722 hCoV-19/England/QEUH-B7F56C/2020\|EPI_ISL_674609\|20 | ···················· | ···················· | ···················· | ···················· | ···················· | ···················· |
| U | 5723 hCoV-19/England/MILK-B8F745/2020\|EPI_ISL_674610\|20 | ···················· | ···················· | ···················· | ···················· | ···················· | ···················· |
| U | 5724 hCoV-19/England/MILK-B8F8D9/2020\|EPI_ISL_674611\|20 | ···················· | ···················· | ···················· | ···················· | ···················· | ···················· |
| U | 5725 hCoV-19/England/QEUH-B808BA/2020\|EPI_ISL_674613\|20 | ···················· | ···················· | ···················· | ···················· | ···················· | ···················· |
| U | 5726 hCoV-19/England/MILK-B8ED17/2020\|EPI_ISL_674614\|20 | ···················· | ···················· | ···················· | ···················· | ···················· | ···················· |
| U | 5727 hCoV-19/England/MILK-B8F94F/2020\|EPI_ISL_674615\|20 | ···················· | ···················· | ···················· | ···················· | ···················· | ···················· |
| U | 5728 hCoV-19/England/MILK-B8ED35/2020\|EPI_ISL_674617\|20 | ···················· | ···················· | ···················· | ···················· | ···················· | ···················· |
| U | 5729 hCoV-19/England/MILK-B8EB2C/2020\|EPI_ISL_674618\|20 | ···················· | ···················· | ···················· | ···················· | ···················· | ···················· |
| U | 5730 hCoV-19/England/MILK-B8DF8B/2020\|EPI_ISL_674619\|20 | ···················· | ···················· | ···················· | ···················· | ·····T·············· | ···················· |
| U | 5731 hCoV-19/England/MILK-B8EBC2/2020\|EPI_ISL_674620\|20 | ···················· | ···················· | ···················· | ···················· | ···················· | ···················· |
| U | 5732 hCoV-19/England/MILK-B8ED44/2020\|EPI_ISL_674621\|20 | ···················· | ···················· | ···················· | ···················· | ···················· | ···················· |
| U | 5733 hCoV-19/England/MILK-B8F6C0/2020\|EPI_ISL_674622\|20 | ···················· | ·A·················· | ···················· | ···················· | ···················· | ··A················· |
| U | 5734 hCoV-19/England/MILK-B8DE06/2020\|EPI_ISL_674623\|20 | ···················· | ···················· | ···················· | ···················· | ···················· | ···················· |
| U | 5735 hCoV-19/England/MILK-B8F675/2020\|EPI_ISL_674624\|20 | ···················· | ···················· | ···················· | ···················· | ···················· | ···················· |
| U | 5736 hCoV-19/England/QEUH-B7CEBF/2020\|EPI_ISL_674625\|20 | ···················· | ···················· | ···················· | ···················· | ···················· | ···················· |
| U | 5737 hCoV-19/England/MILK-B8E45E/2020\|EPI_ISL_674626\|20 | ···················· | ···················· | ···················· | ···················· | ···················· | ···················· |
| U | 5738 hCoV-19/England/MILK-B8F43F/2020\|EPI_ISL_674627\|20 | ···················· | ···················· | ···················· | ···················· | ···················· | ···················· |
| U | 5739 hCoV-19/England/MILK-B8EA6B/2020\|EPI_ISL_674628\|20 | ···················· | ···················· | ···················· | ···················· | ···················· | ···················· |
| U | 5740 hCoV-19/England/MILK-B8EA89/2020\|EPI_ISL_674631\|20 | ···················· | ···················· | ···················· | ···················· | ···················· | ···················· |
| U | 5741 hCoV-19/England/MILK-B8FA2E/2020\|EPI_ISL_674688\|20 | ···················· | ···················· | ···················· | ···················· | ···················· | ···················· |
| U | 5742 hCoV-19/England/MILK-B8ECED/2020\|EPI_ISL_674689\|20 | ···················· | ···················· | ···················· | ···················· | ···················· | ···················· |
| U | 5743 hCoV-19/England/MILK-B8DC0C/2020\|EPI_ISL_674695\|20 | ···················· | ···················· | ···················· | ···················· | ···················· | ···················· |
| U | 5744 hCoV-19/England/MILK-B8F4B7/2020\|EPI_ISL_674697\|20 | ···················· | ·A·······A·········· | ···················· | ···················· | ···················· | ···················· |
| U | 5745 hCoV-19/England/QEUH-B8096C/2020\|EPI_ISL_674698\|20 | ···················· | ···················· | ···················· | ···················· | ···················· | ···················· |
| U | 5746 hCoV-19/England/MILK-B8FA1F/2020\|EPI_ISL_674699\|20 | ···················· | ···················· | ···················· | ···················· | ···················· | ···················· |
| U | 5747 hCoV-19/England/MILK-B8EF02/2020\|EPI_ISL_675170\|20 | ···················· | ···················· | ···················· | ···················· | ···················· | ···················· |
| U | 5748 hCoV-19/England/ALDP-B83A93/2020\|EPI_ISL_675171\|20 | ···················· | ···················· | ···················· | ···················· | ···················· | ···················· |
| U | 5749 hCoV-19/England/ALDP-B82A1C/2020\|EPI_ISL_675174\|20 | ···················· | ···················· | ···················· | ···················· | ···················· | ···················· |
| U | 5750 hCoV-19/England/ALDP-B83C7F/2020\|EPI_ISL_675175\|20 | ···················· | ···················· | ···················· | ···················· | ···················· | ···················· |
| U | 5751 hCoV-19/England/ALDP-B8424F/2020\|EPI_ISL_675176\|20 | ···················· | ···················· | ···················· | ···················· | ···················· | ···················· |
| U | 5752 hCoV-19/England/ALDP-B8418E/2020\|EPI_ISL_675177\|20 | ···················· | ···················· | ···················· | ···················· | ···················· | ··A················· |
| U | 5753 hCoV-19/England/MILK-B8F11A/2020\|EPI_ISL_675178\|20 | ···················· | ···················· | ···················· | ···················· | ···T················ | ···················· |
| U | 5754 hCoV-19/England/MILK-B8EE6F/2020\|EPI_ISL_675179\|20 | ···················· | ···················· | ···················· | ···················· | ···················· | ···················· |
| U | 5755 hCoV-19/England/ALDP-B84616/2020\|EPI_ISL_675181\|20 | ···················· | ···················· | ···················· | ···················· | ···················· | ···················· |
| U | 5756 hCoV-19/England/ALDP-B83714/2020\|EPI_ISL_675182\|20 | ···················· | ···················· | ···················· | ···················· | ···················· | ···················· |
| U | 5757 hCoV-19/England/MILK-B8EF11/2020\|EPI_ISL_675183\|20 | ···················· | ···················· | ···················· | ···················· | ···················· | ···················· |
| U | 5758 hCoV-19/England/ALDP-B856F7/2020\|EPI_ISL_675184\|20 | ···················· | ···················· | ···················· | ···················· | ···················· | ···················· |
| U | 5759 hCoV-19/England/MILK-B8F1DE/2020\|EPI_ISL_675185\|20 | ···················· | ···················· | ···················· | ···················· | ···················· | ···················· |

| | | | | | | | |
|---|---|---|---|---|---|---|---|
| U | 5760 hCoV-19/England/ALDP-B84722/2020\|EPI_ISL_675187\|20 | ···················· | ···················· | ····················· | ···················· | ···················· | ···················· |
| U | 5761 hCoV-19/England/ALDP-B838B7/2020\|EPI_ISL_675189\|20 | ···················· | ···················· | ····················· | ···················· | ···················· | ···················· |
| U | 5762 hCoV-19/England/ALDP-B84018/2020\|EPI_ISL_675190\|20 | ···················· | ···················· | ····················· | ···················· | ···················· | ···················· |
| U | 5763 hCoV-19/England/ALDP-B829C4/2020\|EPI_ISL_675191\|20 | ···················· | ···················· | ····················· | ···················· | ···················· | ···················· |
| U | 5764 hCoV-19/England/ALDP-B83A84/2020\|EPI_ISL_675192\|20 | ···················· | ···················· | ····················· | ···················· | ···················· | ···················· |
| U | 5765 hCoV-19/England/ALDP-B82F68/2020\|EPI_ISL_675193\|20 | ···················· | ······R············· | ····················· | ···················· | ···················· | ···················· |
| U | 5766 hCoV-19/England/ALDP-B829F1/2020\|EPI_ISL_675194\|20 | ···················· | ···················· | ····················· | ···················· | ···················· | ···················· |
| U | 5767 hCoV-19/England/ALDP-B836DB/2020\|EPI_ISL_675195\|20 | ···················· | ···················· | ····················· | ···················· | ···················· | ···················· |
| U | 5768 hCoV-19/England/ALDP-B83A48/2020\|EPI_ISL_675196\|20 | ···················· | ···················· | ····················· | ···················· | ···················· | ···················· |
| U | 5769 hCoV-19/England/ALDP-B82BDD/2020\|EPI_ISL_675197\|20 | ···················· | ···················· | ····················· | ···················· | ···················· | ···················· |
| U | 5770 hCoV-19/England/ALDP-B837AB/2020\|EPI_ISL_675198\|20 | ···················· | ···················· | ····················· | ···················· | ···················· | ···················· |
| U | 5771 hCoV-19/England/MILK-B8C5B7/2020\|EPI_ISL_675900\|20 | ···················· | ···················· | ····················· | ···················· | ···················· | ···················· |
| U | 5772 hCoV-19/England/ALDP-B84C32/2020\|EPI_ISL_675901\|20 | ···················· | ···················· | ····················· | ···················· | ···················· | ···················· |
| U | 5773 hCoV-19/England/ALDP-B84D4E/2020\|EPI_ISL_675902\|20 | ···················· | ···················· | ····················· | ···················· | ···················· | ···················· |
| U | 5774 hCoV-19/England/MILK-B88BF5/2020\|EPI_ISL_675904\|20 | ···················· | ···················· | ····················· | ···················· | ···················· | ···················· |
| U | 5775 hCoV-19/England/MILK-B89541/2020\|EPI_ISL_675905\|20 | ···················· | ···················· | ····················· | ···················· | ···················· | ···················· |
| U | 5776 hCoV-19/England/MILK-B8C933/2020\|EPI_ISL_675907\|20 | ···················· | ···················· | ····················· | ···················· | ···················· | ···················· |
| U | 5777 hCoV-19/England/ALDP-B84E5A/2020\|EPI_ISL_675908\|20 | ···················· | ···················· | ····················· | ···················· | ···················· | ···················· |
| U | 5778 hCoV-19/England/MILK-B897E1/2020\|EPI_ISL_675909\|20 | ···················· | ···················· | ····················· | ···················· | ···················· | ···················· |
| U | 5779 hCoV-19/England/MILK-B88C3E/2020\|EPI_ISL_675910\|20 | ···················· | ···················· | ····················· | ···················· | ······T············· | ···················· |
| U | 5780 hCoV-19/England/MILK-B8C7A2/2020\|EPI_ISL_675911\|20 | ···················· | ···················· | ····················· | ···················· | ···················· | ···················· |
| U | 5781 hCoV-19/England/MILK-B88B6E/2020\|EPI_ISL_675912\|20 | ···················· | ···················· | ····················· | ···················· | ···················· | ···················· |
| U | 5782 hCoV-19/England/ALDP-B84889/2020\|EPI_ISL_675913\|20 | ···················· | ···················· | ····················· | ···················· | ···················· | ···················· |
| U | 5783 hCoV-19/England/MILK-B88C5C/2020\|EPI_ISL_675914\|20 | ···················· | ···················· | ····················· | ···················· | ···················· | ···················· |
| U | 5784 hCoV-19/England/MILK-B89A51/2020\|EPI_ISL_675915\|20 | ···················· | ···················· | ····················· | ···················· | ······T············· | ···················· |
| U | 5785 hCoV-19/England/ALDP-B84E1E/2020\|EPI_ISL_675916\|20 | ···················· | ···················· | ····················· | ···················· | ···················· | ···················· |
| U | 5786 hCoV-19/England/MILK-B8C951/2020\|EPI_ISL_675918\|20 | ···················· | ···················· | ····················· | ···················· | ···················· | ···················· |
| U | 5787 hCoV-19/England/MILK-B8958D/2020\|EPI_ISL_675919\|20 | ···················· | ···················· | ····················· | ···················· | ······T············· | ···················· |
| U | 5788 hCoV-19/England/ALDP-B850CC/2020\|EPI_ISL_675920\|20 | ···················· | ···················· | ····················· | ···················· | ···················· | ···················· |
| U | 5789 hCoV-19/England/MILK-B88B22/2020\|EPI_ISL_675921\|20 | ···················· | ···················· | ····················· | ···················· | ······T············· | ···················· |
| U | 5790 hCoV-19/England/MILK-B8974B/2020\|EPI_ISL_675922\|20 | ···················· | ···················· | ····················· | ···················· | ···················· | ···················· |
| U | 5791 hCoV-19/England/ALDP-B8486B/2020\|EPI_ISL_675923\|20 | ···················· | ···················· | ····················· | ···················· | ···················· | ···················· |
| U | 5792 hCoV-19/England/ALDP-B8509F/2020\|EPI_ISL_675924\|20 | ···················· | ···················· | ····················· | ···················· | ···················· | ···················· |
| U | 5793 hCoV-19/England/ALDP-B85A46/2020\|EPI_ISL_675925\|20 | ···················· | ···················· | ····················· | ···················· | ···················· | ···················· |
| U | 5794 hCoV-19/England/MILK-B889CE/2020\|EPI_ISL_675926\|20 | ···················· | ···················· | ····················· | ···················· | ···················· | ···················· |
| U | 5795 hCoV-19/England/MILK-B8CA5E/2020\|EPI_ISL_675927\|20 | ···················· | ···················· | ····················· | ···················· | ···················· | ···················· |
| U | 5796 hCoV-19/England/ALDP-B859FE/2020\|EPI_ISL_675929\|20 | ···················· | ···················· | ····················· | ···················· | ···················· | ···················· |
| U | 5797 hCoV-19/England/ALDP-B84D6C/2020\|EPI_ISL_675930\|20 | ···················· | ···················· | ····················· | ···················· | ···················· | ···················· |
| U | 5798 hCoV-19/England/MILK-B8C8AF/2020\|EPI_ISL_675931\|20 | ···················· | ···················· | ····················· | ···················· | ···················· | ···················· |
| U | 5799 hCoV-19/England/MILK-B8CA12/2020\|EPI_ISL_675933\|20 | ···················· | ···················· | ····················· | ···················· | ···················· | ···················· |
| U | 5800 hCoV-19/England/ALDP-B848B6/2020\|EPI_ISL_675934\|20 | ···················· | ···················· | ····················· | ···················· | ···················· | ···················· |
| U | 5801 hCoV-19/England/MILK-B8CA8B/2020\|EPI_ISL_675935\|20 | ···················· | ·A·················· | ····················· | ···················· | ···················· | ···A················ |
| U | 5802 hCoV-19/England/MILK-B8C6C3/2020\|EPI_ISL_675936\|20 | ···················· | ···················· | ····················· | ···················· | ···················· | ···················· |
| U | 5803 hCoV-19/England/ALDP-B8482F/2020\|EPI_ISL_675937\|20 | ···················· | ···················· | ····················· | ···················· | ······T············· | ···················· |
| U | 5804 hCoV-19/England/MILK-B896C6/2020\|EPI_ISL_675938\|20 | ···················· | ···················· | ····················· | ···················· | ···················· | ···················· |
| U | 5805 hCoV-19/England/ALDP-B8491D/2020\|EPI_ISL_675939\|20 | ···················· | ·A·················· | ····················· | ···················· | ···················· | ···A················ |
| U | 5806 hCoV-19/England/MILK-B88A9E/2020\|EPI_ISL_675940\|20 | ···················· | ···················· | ····················· | ···················· | ······T············· | ···················· |
| U | 5807 hCoV-19/England/ALDP-B84BDB/2020\|EPI_ISL_675942\|20 | ···················· | ···················· | ····················· | ···················· | ···················· | ···················· |
| U | 5808 hCoV-19/England/MILK-B88858/2020\|EPI_ISL_675943\|20 | ···················· | ···················· | ····················· | ···················· | ···················· | ···················· |
| U | 5809 hCoV-19/England/ALDP-B85044/2020\|EPI_ISL_675944\|20 | ···················· | ···················· | ····················· | ···················· | ···················· | ···················· |
| U | 5810 hCoV-19/England/MILK-B88937/2020\|EPI_ISL_675946\|20 | ···················· | ···················· | ····················· | ···················· | ···················· | ···················· |
| U | 5811 hCoV-19/England/MILK-B88ADA/2020\|EPI_ISL_675947\|20 | ···················· | ···················· | ····················· | ···················· | ······T············· | ···················· |
| U | 5812 hCoV-19/England/MILK-B88AF8/2020\|EPI_ISL_675949\|20 | ···················· | ···················· | ····················· | ···················· | ···················· | ···················· |
| U | 5813 hCoV-19/England/MILK-B88B13/2020\|EPI_ISL_675950\|20 | ···················· | ···················· | ····················· | ···················· | ···················· | ···················· |
| U | 5814 hCoV-19/England/ALDP-B84DD5/2020\|EPI_ISL_675952\|20 | ···················· | ···················· | ····················· | ···················· | ···················· | ···················· |
| U | 5815 hCoV-19/England/MILK-B88B9B/2020\|EPI_ISL_675953\|20 | ···················· | ···················· | ····················· | ···················· | ···················· | ···················· |
| U | 5816 hCoV-19/England/ALDP-B848D4/2020\|EPI_ISL_675954\|20 | ···················· | ···················· | ····················· | ···················· | ···················· | ···················· |
| U | 5817 hCoV-19/England/MILK-B8C890/2020\|EPI_ISL_675957\|20 | ···················· | ···················· | ····················· | ···················· | ······T············· | ···················· |
| U | 5818 hCoV-19/England/MILK-B894CC/2020\|EPI_ISL_675958\|20 | ···················· | ···················· | ····················· | ···················· | ···················· | ···················· |
| U | 5819 hCoV-19/England/MILK-B8C793/2020\|EPI_ISL_675959\|20 | ···················· | ···················· | ····················· | ···················· | ······T············· | ···················· |
| U | 5820 hCoV-19/England/MILK-B89602/2020\|EPI_ISL_675960\|20 | ···················· | ···················· | ····················· | ···················· | ···················· | ···················· |
| U | 5821 hCoV-19/England/MILK-B897A5/2020\|EPI_ISL_675961\|20 | ···················· | ···················· | ····················· | ···················· | ···················· | ···················· |
| U | 5822 hCoV-19/England/ALDP-B8485C/2020\|EPI_ISL_675962\|20 | ···················· | ···················· | ····················· | ···················· | ···················· | ···················· |
| U | 5823 hCoV-19/Scotland/QEUH-B3186D/2020\|EPI_ISL_676415\|2 | ···················· | ···················· | ····················· | ···················· | ···················· | ···················· |
| U | 5824 hCoV-19/Scotland/QEUH-B31BCE/2020\|EPI_ISL_676416\|2 | ···················· | ···················· | ····················· | ···················· | ···················· | ···················· |
| U | 5825 hCoV-19/Scotland/QEUH-B317E8/2020\|EPI_ISL_676417\|2 | ···················· | ···················· | ····················· | ···················· | ···················· | ···················· |
| U | 5826 hCoV-19/Scotland/QEUH-B317CA/2020\|EPI_ISL_676418\|2 | ···················· | ···················· | ····················· | ···················· | ···················· | ···················· |
| U | 5827 hCoV-19/Scotland/QEUH-B31760/2020\|EPI_ISL_676419\|2 | ···················· | ···················· | ····················· | ···················· | ···················· | ···················· |
| U | 5828 hCoV-19/Scotland/QEUH-B3189A/2020\|EPI_ISL_676423\|2 | ···················· | ···················· | ····················· | ···················· | ···················· | ···················· |
| U | 5829 hCoV-19/Scotland/QEUH-B319B5/2020\|EPI_ISL_676424\|2 | ···················· | ···················· | ····················· | ···················· | ···················· | ···················· |
| U | 5830 hCoV-19/Hungary/US-32381w/2020\|EPI_ISL_677746\|2020 | ···················· | ···················· | ····················· | ···················· | ···················· | ···················· |
| U | 5831 hCoV-19/Hungary/US-32488w/2020\|EPI_ISL_677748\|2020 | ···················· | ···················· | ····················· | ···················· | ···················· | ···················· |
| U | 5832 hCoV-19/Hungary/US-32505w/2020\|EPI_ISL_677749\|2020 | ···················· | ···················· | ····················· | ···················· | ···················· | ···················· |
| U | 5833 hCoV-19/Hungary/US-32542w/2020\|EPI_ISL_677754\|2020 | ···················· | ···················· | ····················· | ···················· | ···················· | ···················· |
| U | 5834 hCoV-19/Hungary/US-32545w/2020\|EPI_ISL_677755\|2020 | ···················· | ···················· | ····················· | ···················· | ···················· | ···················· |
| U | 5835 hCoV-19/Saudi Arabia/KAUST-RIYADH1181/2020\|EPI_ISL | ···················· | ·A·················· | ····················· | ···················· | ···················· | ···A················ |
| U | 5836 hCoV-19/Saudi Arabia/KAUST-RIYADH1157/2020\|EPI_ISL | ···················· | ·A·················· | ····················· | ···················· | ···················· | ···A················ |
| U | 5837 hCoV-19/Saudi Arabia/KAUST-RIYADH1163/2020\|EPI_ISL | ···················· | ···················· | ····················· | ···················· | ···················· | ···················· |
| U | 5838 hCoV-19/Saudi Arabia/KAUST-RIYADH1171/2020\|EPI_ISL | ···················· | ···················· | ····················· | ···················· | ···················· | ···················· |
| U | 5839 hCoV-19/Saudi Arabia/KAUST-RIYADH1172/2020\|EPI_ISL | ···················· | ···················· | ····················· | ···················· | ···················· | ···A················ |

| | | | | | | | | |
|---|---|---|---|---|---|---|---|---|
| U | 5840 | hCoV-19/Saudi Arabia/KAUST-RIYADH1174/2020\|EPI_ISL | ···················· | ···················· | ····················· | ···················· | ···················· | ···················· |
| U | 5841 | hCoV-19/Saudi Arabia/KAUST-RIYADH1176/2020\|EPI_ISL | ···················· | ·A·················· | ····················· | ···················· | ···················· | ···A················ |
| U | 5842 | hCoV-19/Saudi Arabia/KAUST-RIYADH1178/2020\|EPI_ISL | ···················· | ···················· | ····················· | ···················· | ···················· | ···················· |
| U | 5843 | hCoV-19/Saudi Arabia/KAUST-RIYADH1180/2020\|EPI_ISL | ···················· | ···················· | ····················· | ···················· | ···················· | ···A················ |
| U | 5844 | hCoV-19/Saudi Arabia/KAUST-RIYADH1191/2020\|EPI_ISL | ···················· | ···················· | ····················· | ···················· | ···················· | ···················· |
| U | 5845 | hCoV-19/Saudi Arabia/KAUST-RIYADH1192/2020\|EPI_ISL | ···················· | ···················· | ····················· | ···················· | ···················· | ···················· |
| U | 5846 | hCoV-19/Scotland/EDB10111/2020\|EPI_ISL_680473\|2020 | ···················· | ···················· | ····················· | ···················· | ···················· | ···················· |
| U | 5848 | hCoV-19/Scotland/EDB10114/2020\|EPI_ISL_680475\|2020 | ···················· | ···················· | ····················· | ···················· | ···················· | ···················· |
| U | 5849 | hCoV-19/Scotland/EDB10115/2020\|EPI_ISL_680476\|2020 | ···················· | ···················· | ····················· | ···················· | ···················· | ···················· |
| U | 5850 | hCoV-19/Scotland/EDB10116/2020\|EPI_ISL_680477\|2020 | ···················· | ···················· | ····················· | ···················· | ···················· | ···················· |
| U | 5851 | hCoV-19/Scotland/EDB10117/2020\|EPI_ISL_680478\|2020 | ···················· | ···················· | ····················· | ···················· | ···················· | ···················· |
| U | 5852 | hCoV-19/Scotland/EDB10119/2020\|EPI_ISL_680480\|2020 | ···················· | ···················· | ····················· | ···················· | ···················· | ···················· |
| U | 5853 | hCoV-19/Scotland/EDB10121/2020\|EPI_ISL_680481\|2020 | ···················· | ···················· | ····················· | ···················· | ···················· | ···················· |
| U | 5854 | hCoV-19/Scotland/EDB10124/2020\|EPI_ISL_680484\|2020 | ···················· | ···················· | ····················· | ···················· | ···················· | ···················· |
| U | 5855 | hCoV-19/Scotland/EDB10125/2020\|EPI_ISL_680485\|2020 | ···················· | ···················· | ····················· | ···················· | ···················· | ···················· |
| U | 5856 | hCoV-19/Scotland/EDB10130/2020\|EPI_ISL_680486\|2020 | ···················· | ···················· | ····················· | ···················· | ···················· | ···················· |
| U | 5858 | hCoV-19/Scotland/EDB10157/2020\|EPI_ISL_680489\|2020 | ···················· | ···················· | ····················· | ···················· | ···················· | ···················· |
| U | 5859 | hCoV-19/Scotland/EDB10164/2020\|EPI_ISL_680490\|2020 | ···················· | ···················· | ····················· | ···················· | ···················· | ···················· |
| U | 5860 | hCoV-19/Scotland/EDB10169/2020\|EPI_ISL_680491\|2020 | ···················· | ···················· | ····················· | ···················· | ···················· | ···················· |
| U | 5861 | hCoV-19/Scotland/EDB10177/2020\|EPI_ISL_680492\|2020 | ···················· | ···················· | ····················· | ···················· | ···················· | ···················· |
| U | 5862 | hCoV-19/Scotland/EDB10180/2020\|EPI_ISL_680493\|2020 | ···················· | ···················· | ····················· | ···················· | ···················· | ···················· |
| U | 5863 | hCoV-19/Scotland/EDB10194/2020\|EPI_ISL_680495\|2020 | ···················· | ···················· | ····················· | ···················· | ···················· | ···················· |
| U | 5864 | hCoV-19/Scotland/EDB10195/2020\|EPI_ISL_680496\|2020 | ···················· | ···················· | ····················· | ···················· | ···················· | ···················· |
| U | 5865 | hCoV-19/Scotland/EDB10196/2020\|EPI_ISL_680497\|2020 | ···················· | ···················· | ····················· | ···················· | ···················· | ···················· |
| U | 5866 | hCoV-19/Scotland/EDB10197/2020\|EPI_ISL_680498\|2020 | ···················· | ···················· | ····················· | ···················· | ···················· | ···················· |
| U | 5867 | hCoV-19/Scotland/EDB10198/2020\|EPI_ISL_680499\|2020 | ···················· | ···················· | ····················· | ···················· | ···················· | ···················· |
| U | 5879 | hCoV-19/USA/UT-UPHL-201218864/2020\|EPI_ISL_684016\| | ···················· | ·A·················· | ····················· | ···················· | ···················· | ···················· |
| U | 5880 | hCoV-19/Bahrain/341036861/2020\|EPI_ISL_684028\|2020 | ···················· | ···················· | ····················· | ···················· | ···················· | ···················· |
| U | 5881 | hCoV-19/Bahrain/341036654/2020\|EPI_ISL_684029\|2020 | ·T·················· | ···················· | ····················· | ···················· | ···················· | ···················· |
| U | 5882 | hCoV-19/Bahrain/341035032/2020\|EPI_ISL_684030\|2020 | ···················· | ·A·················· | ····················· | ···················· | ···················· | ···A················ |
| U | 5883 | hCoV-19/Bahrain/920509111/2020\|EPI_ISL_684031\|2020 | ···················· | ···················· | ····················· | ···················· | ···················· | ···················· |
| U | 5884 | hCoV-19/Bahrain/341033465/2020\|EPI_ISL_684032\|2020 | ···················· | ····A··············· | ····················· | ···················· | ···················· | ···················· |
| U | 5885 | hCoV-19/Bahrain/341033460/2020\|EPI_ISL_684033\|2020 | ···················· | ····A··············· | ····················· | ···················· | ···················· | ···················· |
| U | 5886 | hCoV-19/Bahrain/341017372/2020\|EPI_ISL_684034\|2020 | ···················· | ···················· | ····················· | ···················· | ···················· | ···················· |
| U | 5887 | hCoV-19/Bahrain/341017320/2020\|EPI_ISL_684035\|2020 | ···················· | ···················· | ····················· | ···················· | ·······N············ | ···················· |
| U | 5888 | hCoV-19/Bahrain/250182775/2020\|EPI_ISL_684036\|2020 | ···················· | ···················· | ····················· | ···················· | ···················· | ···················· |
| U | 5889 | hCoV-19/England/CAMC-B99B5B/2020\|EPI_ISL_673268\|20 | ···················· | ···················· | ····················· | ···················· | ···················· | ···················· |
| U | 5890 | hCoV-19/England/CAMC-B995E4/2020\|EPI_ISL_673269\|20 | ···················· | ···················· | ····················· | ···················· | ···················· | ···················· |
| U | 5891 | hCoV-19/England/CAMC-B98F6F/2020\|EPI_ISL_673270\|20 | ···················· | ···················· | ····················· | ···················· | ······T············· | ···················· |
| U | 5892 | hCoV-19/England/CAMC-B98FF6/2020\|EPI_ISL_673271\|20 | ···················· | ···················· | ····················· | ···················· | ···················· | ···················· |
| U | 5893 | hCoV-19/England/CAMC-B9A9AB/2020\|EPI_ISL_673272\|20 | ···················· | ···················· | ····················· | ···················· | ···················· | ···················· |
| U | 5894 | hCoV-19/England/CAMC-B9A0E2/2020\|EPI_ISL_673273\|20 | ···················· | ···················· | ····················· | ···················· | ···················· | ···················· |
| U | 5895 | hCoV-19/England/CAMC-B9960F/2020\|EPI_ISL_673274\|20 | ···················· | ···················· | ····················· | ···················· | ···················· | ···················· |
| U | 5896 | hCoV-19/England/CAMC-B9AFD6/2020\|EPI_ISL_673276\|20 | ···················· | ···················· | ····················· | ···················· | ···················· | ···················· |
| U | 5897 | hCoV-19/England/CAMC-B9A4C8/2020\|EPI_ISL_673277\|20 | ···················· | ···················· | ····················· | ···················· | ···················· | ···················· |
| U | 5898 | hCoV-19/England/CAMC-B99BB5/2020\|EPI_ISL_673278\|20 | ···················· | ···················· | ····················· | ···················· | ···················· | ···················· |
| U | 5899 | hCoV-19/England/CAMC-B98EAE/2020\|EPI_ISL_673279\|20 | ···················· | ···················· | ····················· | ···················· | ···················· | ···················· |
| U | 5900 | hCoV-19/England/CAMC-B9C4A8/2020\|EPI_ISL_673280\|20 | ···················· | ···················· | ····················· | ···················· | ···················· | ···················· |
| U | 5901 | hCoV-19/England/CAMC-B99502/2020\|EPI_ISL_673281\|20 | ···················· | ···················· | ····················· | ···················· | ···················· | ···················· |
| U | 5902 | hCoV-19/England/CAMC-B99890/2020\|EPI_ISL_673282\|20 | ···················· | ···················· | ····················· | ···················· | ···················· | ···················· |
| U | 5903 | hCoV-19/England/CAMC-B99A30/2020\|EPI_ISL_673286\|20 | ···················· | ···················· | ····················· | ···················· | ···················· | ···················· |
| U | 5904 | hCoV-19/England/CAMC-B99678/2020\|EPI_ISL_673287\|20 | ···················· | ···················· | ····················· | ···················· | ···················· | ···················· |
| U | 5905 | hCoV-19/England/CAMC-B9A501/2020\|EPI_ISL_673288\|20 | ···················· | ···················· | ····················· | ···················· | ···················· | ···················· |
| U | 5906 | hCoV-19/England/CAMC-B9A8CC/2020\|EPI_ISL_673289\|20 | ···················· | ···················· | ····················· | ···················· | ···················· | ···················· |
| U | 5907 | hCoV-19/England/CAMC-B99845/2020\|EPI_ISL_673290\|20 | ···················· | ···················· | ····················· | ···················· | ···················· | ···················· |
| U | 5908 | hCoV-19/England/CAMC-B9AFA9/2020\|EPI_ISL_673291\|20 | ···················· | ·A·················· | ····················· | ···················· | ···················· | ···A················ |
| U | 5909 | hCoV-19/England/CAMC-B9961E/2020\|EPI_ISL_673292\|20 | ···················· | ···················· | ····················· | ···················· | ···················· | ···················· |
| U | 5910 | hCoV-19/England/CAMC-B99DBF/2020\|EPI_ISL_673293\|20 | ···················· | ···················· | ····················· | ···················· | ···················· | ···················· |
| U | 5911 | hCoV-19/England/CAMC-B9A01F/2020\|EPI_ISL_673295\|20 | ···················· | ···················· | ····················· | ···················· | ······T············· | ···················· |
| U | 5912 | hCoV-19/England/CAMC-B99326/2020\|EPI_ISL_673296\|20 | ···················· | ···················· | ····················· | ···················· | ···················· | ···················· |
| U | 5913 | hCoV-19/England/CAMC-B9ABF0/2020\|EPI_ISL_673297\|20 | ···················· | ···················· | ····················· | ···················· | ···················· | ···················· |
| U | 5914 | hCoV-19/England/CAMC-B98E80/2020\|EPI_ISL_673298\|20 | ···················· | ···················· | ····················· | ···················· | ···················· | ···················· |
| U | 5915 | hCoV-19/England/CAMC-B99739/2020\|EPI_ISL_673299\|20 | ···················· | ···················· | ····················· | ···················· | ···················· | ···················· |
| U | 5916 | hCoV-19/England/CAMC-B99511/2020\|EPI_ISL_673301\|20 | ···················· | ···················· | ····················· | ···················· | ···················· | ···················· |
| U | 5917 | hCoV-19/England/CAMC-B9AFE5/2020\|EPI_ISL_673302\|20 | ···················· | ···················· | ····················· | ···················· | ···················· | ···················· |
| U | 5918 | hCoV-19/England/CAMC-B9ADBE/2020\|EPI_ISL_673305\|20 | ···················· | ·A·················· | ····················· | ···················· | ···················· | ···A················ |
| U | 5919 | hCoV-19/England/CAMC-B99D64/2020\|EPI_ISL_673306\|20 | ···················· | ···················· | ····················· | ···················· | ···················· | ···················· |
| U | 5920 | hCoV-19/England/CAMC-B98F7E/2020\|EPI_ISL_673308\|20 | ···················· | ···················· | ····················· | ···················· | ···················· | ···················· |
| U | 5921 | hCoV-19/England/CAMC-B99DEC/2020\|EPI_ISL_673309\|20 | ···················· | ···················· | ····················· | ···················· | ···················· | ···················· |
| U | 5922 | hCoV-19/England/CAMC-B9B02D/2020\|EPI_ISL_673310\|20 | ···················· | ···················· | ····················· | ···················· | ···················· | ···················· |
| U | 5923 | hCoV-19/England/CAMC-B997EE/2020\|EPI_ISL_673311\|20 | ···················· | ···················· | ····················· | ···················· | ···················· | ···················· |
| U | 5924 | hCoV-19/England/CAMC-B98B2F/2020\|EPI_ISL_673312\|20 | ···················· | ···················· | ····················· | ···················· | ···················· | ···················· |
| U | 5925 | hCoV-19/England/CAMC-B9A404/2020\|EPI_ISL_673313\|20 | ···················· | ···················· | ····················· | ···················· | ···················· | ···················· |
| U | 5926 | hCoV-19/England/CAMC-B9A06A/2020\|EPI_ISL_673314\|20 | ···················· | ···················· | ····················· | ···················· | ···················· | ···················· |
| U | 5927 | hCoV-19/England/CAMC-B98F8D/2020\|EPI_ISL_673315\|20 | ···················· | ···················· | ····················· | ···················· | ···················· | ···················· |
| U | 5928 | hCoV-19/England/CAMC-B99766/2020\|EPI_ISL_673316\|20 | ···················· | ···················· | ····················· | ···················· | ···················· | ···················· |
| U | 5929 | hCoV-19/England/CAMC-B991C2/2020\|EPI_ISL_673318\|20 | ···················· | ·A·················· | ····················· | ···················· | ···················· | ···A················ |
| U | 5930 | hCoV-19/England/CAMC-B9A9D8/2020\|EPI_ISL_673320\|20 | ···················· | ···················· | ····················· | ···················· | ···················· | ···················· |
| U | 5931 | hCoV-19/England/CAMC-B9C61B/2020\|EPI_ISL_673321\|20 | ···················· | ···················· | ····················· | ···················· | ···················· | ···················· |
| U | 5932 | hCoV-19/England/CAMC-B99C2B/2020\|EPI_ISL_673324\|20 | ···················· | ···················· | ····················· | ···················· | ···················· | ···················· |

| | | | | | | | |
|---|---|---|---|---|---|---|---|
| U | 5933 hCoV-19/England/CAMC-B9921A/2020\|EPI_ISL_673326\|20 | ···················· | ···················· | ····················· | ···················· | ···················· | ···················· |
| U | 5934 hCoV-19/England/CAMC-B9A39E/2020\|EPI_ISL_673328\|20 | ···················· | ···················· | ····················· | ···················· | ···················· | ···················· |
| U | 5935 hCoV-19/England/CAMC-B99AA9/2020\|EPI_ISL_673329\|20 | ···················· | ···················· | ····················· | ···················· | ···················· | ···················· |
| U | 5936 hCoV-19/England/CAMC-B99775/2020\|EPI_ISL_673330\|20 | ···················· | ···················· | ····················· | ···················· | ···················· | ···················· |
| U | 5937 hCoV-19/England/CAMC-B9A3CB/2020\|EPI_ISL_673332\|20 | ···················· | ···················· | ····················· | ···················· | ···················· | ···················· |
| U | 5938 hCoV-19/England/CAMC-B9A2DD/2020\|EPI_ISL_673333\|20 | ···················· | ···················· | ····················· | ···················· | ···················· | ···················· |
| U | 5939 hCoV-19/England/CAMC-B99229/2020\|EPI_ISL_673334\|20 | ···················· | ···················· | ····················· | ···················· | ···················· | ···················· |
| U | 5940 hCoV-19/England/CAMC-B98EDB/2020\|EPI_ISL_673335\|20 | ···················· | ···················· | ····················· | ···················· | ···················· | ···················· |
| U | 5941 hCoV-19/England/CAMC-B990E3/2020\|EPI_ISL_673337\|20 | ···················· | ···················· | ····················· | ···················· | ···················· | ···················· |
| U | 5942 hCoV-19/England/CAMC-B99D19/2020\|EPI_ISL_673338\|20 | ···················· | ···················· | ····················· | ···················· | ···················· | ···················· |
| U | 5943 hCoV-19/England/CAMC-B99CEF/2020\|EPI_ISL_673340\|20 | ···················· | ···················· | ····················· | ···················· | ···················· | ···················· |
| U | 5944 hCoV-19/England/CAMC-B99A21/2020\|EPI_ISL_673341\|20 | ···················· | ···················· | ····················· | ···················· | ···················· | ···················· |
| U | 5945 hCoV-19/England/CAMC-B9C52D/2020\|EPI_ISL_673344\|20 | ···················· | ···················· | ····················· | ···················· | ···················· | ···················· |
| U | 5946 hCoV-19/England/CAMC-B9A9C9/2020\|EPI_ISL_673346\|20 | ···················· | ···················· | ····················· | ···················· | ······T············· | ···················· |
| U | 5947 hCoV-19/England/CAMC-B990B6/2020\|EPI_ISL_673347\|20 | ········T··········· | ···················· | ····················· | ···················· | ···················· | ···················· |
| U | 5948 hCoV-19/England/CAMC-B9C54B/2020\|EPI_ISL_673348\|20 | ···················· | ···················· | ····················· | ···················· | ······T············· | ···················· |
| U | 5949 hCoV-19/England/CAMC-B98F50/2020\|EPI_ISL_673349\|20 | ···················· | ···················· | ····················· | ···················· | ······T············· | ···················· |
| U | 5950 hCoV-19/England/CAMC-B99CB2/2020\|EPI_ISL_673350\|20 | ···················· | ···················· | ····················· | ···················· | ···················· | ···················· |
| U | 5951 hCoV-19/England/CAMC-B996A5/2020\|EPI_ISL_673351\|20 | ···················· | ···················· | ····················· | ···················· | ···················· | ···················· |
| U | 5952 hCoV-19/England/CAMC-B9A0C4/2020\|EPI_ISL_673352\|20 | ···················· | ···················· | ····················· | ···················· | ···················· | ···················· |
| U | 5953 hCoV-19/England/CAMC-B99FC8/2020\|EPI_ISL_673353\|20 | ···················· | ···················· | ····················· | ···················· | ···················· | ···················· |
| U | 5954 hCoV-19/England/CAMC-B98F05/2020\|EPI_ISL_673354\|20 | ···················· | ···················· | ····················· | ···················· | ···················· | ···················· |
| U | 5955 hCoV-19/England/CAMC-B9A0D3/2020\|EPI_ISL_673360\|20 | ···················· | ···················· | ····················· | ···················· | ···················· | ···················· |
| U | 5956 hCoV-19/England/CAMC-B990C5/2020\|EPI_ISL_673361\|20 | ···················· | ···················· | ····················· | ···················· | ···················· | ···················· |
| U | 5957 hCoV-19/England/CAMC-B9A48C/2020\|EPI_ISL_673362\|20 | ···················· | ···················· | ····················· | ···················· | ···················· | ···················· |
| U | 5958 hCoV-19/England/CAMC-B99C67/2020\|EPI_ISL_673363\|20 | ···················· | ···················· | ····················· | ···················· | ···················· | ···················· |
| U | 5959 hCoV-19/England/CAMC-B98D83/2020\|EPI_ISL_673364\|20 | ···················· | ···················· | ····················· | ···················· | ···················· | ···················· |
| U | 5960 hCoV-19/England/MILK-B8E0E1/2020\|EPI_ISL_674599\|20 | ···················· | ···················· | ····················· | ···················· | ···················· | ···················· |
| U | 5961 hCoV-19/England/MILK-B8F912/2020\|EPI_ISL_674632\|20 | ···················· | ···················· | ····················· | ···················· | ···················· | ···················· |
| U | 5962 hCoV-19/England/MILK-B8F87F/2020\|EPI_ISL_674634\|20 | ···················· | ···················· | ····················· | ···················· | ···················· | ···················· |
| U | 5963 hCoV-19/England/QEUH-B7F793/2020\|EPI_ISL_674635\|20 | ···················· | ···················· | ····················· | ···················· | ···················· | ···················· |
| U | 5964 hCoV-19/England/MILK-B8F754/2020\|EPI_ISL_674636\|20 | ···················· | ···················· | ····················· | ···················· | ···················· | ···················· |
| U | 5965 hCoV-19/England/MILK-B8EDDB/2020\|EPI_ISL_674638\|20 | ···················· | ···················· | ····················· | ···················· | ···················· | ···················· |
| U | 5966 hCoV-19/England/MILK-B8F8F7/2020\|EPI_ISL_674639\|20 | ···················· | ···················· | ····················· | ···················· | ···················· | ···················· |
| U | 5967 hCoV-19/England/MILK-B8EC1A/2020\|EPI_ISL_674640\|20 | ···················· | ···················· | ····················· | ···················· | ···················· | ···················· |
| U | 5968 hCoV-19/England/MILK-B8E139/2020\|EPI_ISL_674641\|20 | ···················· | ···················· | ····················· | ···················· | ···················· | ···················· |
| U | 5969 hCoV-19/England/MILK-B8E49A/2020\|EPI_ISL_674642\|20 | ···················· | ···················· | ····················· | ···················· | ···················· | ···················· |
| U | 5970 hCoV-19/England/MILK-B8E078/2020\|EPI_ISL_674643\|20 | ···················· | ···················· | ····················· | ···················· | ···················· | ···················· |
| U | 5971 hCoV-19/England/MILK-B8E47C/2020\|EPI_ISL_674644\|20 | ···················· | ···················· | ····················· | ···················· | ···················· | ···················· |
| U | 5972 hCoV-19/England/MILK-B8F5F0/2020\|EPI_ISL_674646\|20 | ···················· | ···················· | ····················· | ···················· | ···················· | ···················· |
| U | 5973 hCoV-19/England/MILK-B8ECCF/2020\|EPI_ISL_674647\|20 | ···················· | ···················· | ····················· | ···················· | ···················· | ···················· |
| U | 5974 hCoV-19/England/MILK-B8DFE5/2020\|EPI_ISL_674648\|20 | ···················· | ·A·················· | ····················· | ···················· | ···················· | ··A················· |
| U | 5975 hCoV-19/England/MILK-B8E087/2020\|EPI_ISL_674649\|20 | ···················· | ···················· | ····················· | ···················· | ···················· | ···················· |
| U | 5976 hCoV-19/England/MILK-B8ECA1/2020\|EPI_ISL_674650\|20 | ···················· | ···················· | ····················· | ···················· | ···················· | ···················· |
| U | 5977 hCoV-19/England/MILK-B8EDAE/2020\|EPI_ISL_674651\|20 | ···················· | ···················· | ····················· | ···················· | ···················· | ···················· |
| U | 5978 hCoV-19/England/QEUH-B7CDFE/2020\|EPI_ISL_674652\|20 | ···················· | ···················· | ····················· | ···················· | ···················· | ···················· |
| U | 5979 hCoV-19/England/MILK-B8F718/2020\|EPI_ISL_674654\|20 | ···················· | ···················· | ····················· | ···················· | ···················· | ···················· |
| U | 5980 hCoV-19/England/QEUH-B7F599/2020\|EPI_ISL_674655\|20 | ···················· | ···················· | ····················· | ···················· | ···················· | ···················· |
| U | 5981 hCoV-19/England/MILK-B8E913/2020\|EPI_ISL_674656\|20 | ···················· | ···················· | ····················· | ···················· | ···················· | ···················· |
| U | 5982 hCoV-19/England/QEUH-B7F4F6/2020\|EPI_ISL_674657\|20 | ···················· | ···················· | ····················· | ···················· | ···················· | ···················· |
| U | 5983 hCoV-19/England/QEUH-B7F502/2020\|EPI_ISL_674658\|20 | ···················· | ···················· | ····················· | ···················· | ···················· | ···················· |
| U | 5984 hCoV-19/England/MILK-B8E931/2020\|EPI_ISL_674660\|20 | ···················· | ···················· | ····················· | ···················· | ···················· | ···················· |
| U | 5985 hCoV-19/England/QEUH-B7D689/2020\|EPI_ISL_674661\|20 | ···················· | ···················· | ····················· | ···················· | ···················· | ···················· |
| U | 5986 hCoV-19/England/QEUH-B80911/2020\|EPI_ISL_674662\|20 | ···················· | ···················· | ····················· | ···················· | ···················· | ···················· |
| U | 5987 hCoV-19/England/MILK-B8EA5C/2020\|EPI_ISL_674663\|20 | ···················· | ···················· | ····················· | ···················· | ······T············· | ···················· |
| U | 5988 hCoV-19/England/MILK-B8DE51/2020\|EPI_ISL_674664\|20 | ···················· | ···················· | ····················· | ···················· | ···················· | ···················· |
| U | 5989 hCoV-19/England/MILK-B8DDDC/2020\|EPI_ISL_674666\|20 | ···················· | ·A·················· | ····················· | ···················· | ···················· | ··A················· |
| U | 5990 hCoV-19/England/QEUH-B7CE19/2020\|EPI_ISL_674667\|20 | ···················· | ···················· | ····················· | ···················· | ···················· | ···················· |
| U | 5991 hCoV-19/England/MILK-B8E12A/2020\|EPI_ISL_674668\|20 | ···················· | ···················· | ····················· | ···················· | ···················· | ···················· |
| U | 5992 hCoV-19/England/MILK-B8DD81/2020\|EPI_ISL_674669\|20 | ···················· | ···················· | ····················· | ···················· | ···················· | ···················· |
| U | 5993 hCoV-19/England/MILK-B8EA4D/2020\|EPI_ISL_674673\|20 | ···················· | ···················· | ····················· | ···················· | ···················· | ···················· |
| U | 5994 hCoV-19/England/MILK-B8DD36/2020\|EPI_ISL_674675\|20 | ···················· | ···················· | ····················· | ···················· | ···················· | ···················· |
| U | 5995 hCoV-19/England/MILK-B8EDCC/2020\|EPI_ISL_674676\|20 | ···················· | ···················· | ····················· | ···················· | ···················· | ···················· |
| U | 5996 hCoV-19/England/QEUH-B7D62F/2020\|EPI_ISL_674678\|20 | ···················· | ···················· | ····················· | ···················· | ···················· | ···················· |
| U | 5997 hCoV-19/England/MILK-B8E0B4/2020\|EPI_ISL_674679\|20 | ···················· | ···················· | ····················· | ···················· | ···················· | ···················· |
| U | 5998 hCoV-19/England/MILK-B8EAF2/2020\|EPI_ISL_674680\|20 | ···················· | ···················· | ····················· | ···················· | ···················· | ···················· |
| U | 5999 hCoV-19/England/MILK-B8EA3E/2020\|EPI_ISL_674681\|20 | ···················· | ···················· | ····················· | ···················· | ···················· | ···················· |
| U | 6000 hCoV-19/England/MILK-B8DE24/2020\|EPI_ISL_674682\|20 | ···················· | ·A·················· | ····················· | ···················· | ···················· | ··A················· |
| U | 6001 hCoV-19/England/MILK-B8DC93/2020\|EPI_ISL_674683\|20 | ···················· | ···················· | ····················· | ···················· | ···················· | ···················· |
| U | 6002 hCoV-19/England/MILK-B8E51F/2020\|EPI_ISL_674684\|20 | ···················· | ···················· | ····················· | ···················· | ···················· | ···················· |
| U | 6003 hCoV-19/England/QEUH-B7F818/2020\|EPI_ISL_674685\|20 | ···················· | ···················· | ····················· | ···················· | ···················· | ···················· |
| U | 6004 hCoV-19/England/QEUH-B808E7/2020\|EPI_ISL_674690\|20 | ···················· | ···················· | ····················· | ···················· | ···················· | ···················· |
| U | 6005 hCoV-19/England/QEUH-B7F7A2/2020\|EPI_ISL_674691\|20 | ···················· | ·A·················· | ····················· | ···················· | ···················· | ··A················· |
| U | 6006 hCoV-19/England/QEUH-B7F49C/2020\|EPI_ISL_674692\|20 | ···················· | ···················· | ····················· | ···················· | ···················· | ···················· |
| U | 6007 hCoV-19/England/MILK-B8EC0B/2020\|EPI_ISL_674693\|20 | ···················· | ···················· | ····················· | ···················· | ···················· | ···················· |
| U | 6008 hCoV-19/England/MILK-B8EC92/2020\|EPI_ISL_674694\|20 | ···················· | ···················· | ····················· | ···················· | ···················· | ···················· |
| U | 6009 hCoV-19/England/MILK-B8C818/2020\|EPI_ISL_675963\|20 | ···················· | ···················· | ····················· | ···················· | ···················· | ···················· |
| U | 6010 hCoV-19/England/MILK-B8C71B/2020\|EPI_ISL_675964\|20 | ···················· | ···················· | ····················· | ···················· | ···················· | ···················· |
| U | 6011 hCoV-19/England/ALDP-B850AE/2020\|EPI_ISL_675965\|20 | ···················· | ···················· | ····················· | ···················· | ···················· | ···················· |
| U | 6012 hCoV-19/England/ALDP-B84B71/2020\|EPI_ISL_675968\|20 | ···················· | ···················· | ····················· | ···················· | ···················· | ···················· |

| | | | | | | | |
|---|---|---|---|---|---|---|---|
| U | 6013 hCoV-19/England/MILK-B88C01/2020\|EPI_ISL_675971\|20 | ···················· | ···················· | ···················· | ···················· | ···················· | ···················· |
| U | 6014 hCoV-19/England/ALDP-B84C50/2020\|EPI_ISL_675974\|20 | ···················· | ···················· | ···················· | ···················· | ···················· | ···················· |
| U | 6015 hCoV-19/England/MILK-B8C9BB/2020\|EPI_ISL_675975\|20 | ···················· | ···················· | ···················· | ···················· | ···················· | ···················· |
| U | 6016 hCoV-19/England/ALDP-B847F5/2020\|EPI_ISL_675976\|20 | ···················· | ·A·················· | ···················· | ···················· | ···················· | ···A················ |
| U | 6017 hCoV-19/England/ALDP-B84898/2020\|EPI_ISL_675980\|20 | ···················· | ···················· | ···················· | ···················· | ···················· | ···················· |
| U | 6018 hCoV-19/England/MILK-B89611/2020\|EPI_ISL_675981\|20 | ···················· | ···················· | ···················· | ···················· | ···················· | ···················· |
| U | 6019 hCoV-19/England/ALDP-B84EE1/2020\|EPI_ISL_675982\|20 | ···················· | ···················· | ···················· | ···················· | ···················· | ···················· |
| U | 6020 hCoV-19/England/MILK-B888FE/2020\|EPI_ISL_675983\|20 | ···················· | ···················· | ···················· | ···················· | ···················· | ···················· |
| U | 6021 hCoV-19/England/MILK-B88ACB/2020\|EPI_ISL_675984\|20 | ···················· | ···················· | ···················· | ···················· | ···················· | ···················· |
| U | 6022 hCoV-19/England/MILK-B89523/2020\|EPI_ISL_675985\|20 | ···················· | ···················· | ···················· | ···················· | ···················· | ···················· |
| U | 6023 hCoV-19/England/ALDP-B84E0F/2020\|EPI_ISL_675986\|20 | ···················· | ···················· | ···················· | ···················· | ···················· | ···················· |
| U | 6024 hCoV-19/England/ALDP-B85A37/2020\|EPI_ISL_675987\|20 | ···················· | ···················· | ···················· | ···················· | ···················· | ···················· |
| U | 6025 hCoV-19/England/ALDP-B848E3/2020\|EPI_ISL_675988\|20 | ···················· | ·A·················· | ···················· | ···················· | ···················· | ···A················ |
| U | 6026 hCoV-19/England/MILK-B896F3/2020\|EPI_ISL_675990\|20 | ···················· | ···················· | ···················· | ···················· | ···················· | ···················· |
| U | 6027 hCoV-19/England/MILK-B8C61E/2020\|EPI_ISL_675991\|20 | ···················· | ···················· | ···················· | ···················· | ······T············· | ···················· |
| U | 6028 hCoV-19/England/ALDP-B84E87/2020\|EPI_ISL_675993\|20 | ···················· | ···················· | ···················· | ···················· | ···················· | ···················· |
| U | 6029 hCoV-19/England/MILK-B8C809/2020\|EPI_ISL_675996\|20 | ···················· | ···················· | ···················· | ···················· | ···················· | ···················· |
| U | 6030 hCoV-19/England/MILK-B8C99D/2020\|EPI_ISL_675997\|20 | ···················· | ···················· | ···················· | ···················· | ···················· | ···················· |
| U | 6031 hCoV-19/England/MILK-B8C854/2020\|EPI_ISL_675998\|20 | ···················· | ···················· | ···················· | ···················· | ···················· | ···················· |
| U | 6032 hCoV-19/England/MILK-B887E2/2020\|EPI_ISL_675999\|20 | ···················· | ···················· | ···················· | ···················· | ···················· | ···················· |
| U | 6033 hCoV-19/Saudi Arabia/KAUST-MAKKAH1557/2020\|EPI_ISL | ···················· | ···················· | ···················· | ···················· | ···················· | ···················· |
| U | 6034 hCoV-19/Saudi Arabia/KAUST-MAKKAH1592/2020\|EPI_ISL | ···················· | ···················· | ···················· | ···················· | ···················· | ···················· |
| U | 6035 hCoV-19/Saudi Arabia/KAUST-MAKKAH1595/2020\|EPI_ISL | ···················· | ···················· | ···················· | ···················· | ···················· | ···················· |
| U | 6036 hCoV-19/Saudi Arabia/KAUST-MAKKAH1596/2020\|EPI_ISL | ···················· | ···················· | ···················· | ···················· | ···················· | ···················· |
| U | 6037 hCoV-19/Saudi Arabia/KAUST-MAKKAH1602/2020\|EPI_ISL | ···················· | ···················· | ···················· | ···················· | ···················· | ···················· |
| U | 6038 hCoV-19/Saudi Arabia/KAUST-MAKKAH1604/2020\|EPI_ISL | ···················· | ···················· | ···················· | ···················· | ···················· | ···················· |
| U | 6039 hCoV-19/Saudi Arabia/KAUST-MAKKAH1605/2020\|EPI_ISL | ···················· | ···················· | ···················· | ···················· | ···················· | ···················· |
| U | 6040 hCoV-19/Saudi Arabia/KAUST-MAKKAH1611/2020\|EPI_ISL | ···················· | ···················· | ···················· | ···················· | ···················· | ···················· |
| U | 6041 hCoV-19/Saudi Arabia/KAUST-MAKKAH1617/2020\|EPI_ISL | ···················· | ···················· | ···················· | ···················· | ···················· | ···················· |
| U | 6042 hCoV-19/Saudi Arabia/KAUST-MAKKAH162/2020\|EPI_ISL_ | ···················· | ···················· | ···················· | ···················· | ···················· | ···················· |
| U | 6043 hCoV-19/Saudi Arabia/KAUST-MAKKAH1621/2020\|EPI_ISL | ···················· | ···················· | ···················· | ···················· | ···················· | ···················· |
| U | 6044 hCoV-19/Saudi Arabia/KAUST-MAKKAH1623/2020\|EPI_ISL | ···················· | ···················· | ···················· | ···················· | ···················· | ···················· |
| U | 6045 hCoV-19/Saudi Arabia/KAUST-MAKKAH1626/2020\|EPI_ISL | ···················· | ···················· | ···················· | ···················· | ···················· | ···················· |
| U | 6046 hCoV-19/Saudi Arabia/KAUST-MAKKAH1630/2020\|EPI_ISL | ···················· | ·A·················· | ···················· | ···················· | ···················· | ···A················ |
| U | 6047 hCoV-19/Saudi Arabia/KAUST-MAKKAH1631/2020\|EPI_ISL | ···················· | ···················· | ···················· | ···················· | ···················· | ···················· |
| U | 6048 hCoV-19/Saudi Arabia/KAUST-MAKKAH1632/2020\|EPI_ISL | ···················· | ···················· | ···················· | ···················· | ···················· | ···················· |
| U | 6049 hCoV-19/Saudi Arabia/KAUST-MAKKAH1635/2020\|EPI_ISL | ···················· | ·A·················· | ···················· | ···················· | ···················· | ···················· |
| U | 6050 hCoV-19/Saudi Arabia/KAUST-MAKKAH167/2020\|EPI_ISL_ | ···················· | ···················· | ···················· | ···················· | ···················· | ···················· |
| U | 6051 hCoV-19/Saudi Arabia/KAUST-MAKKAH172/2020\|EPI_ISL_ | ···················· | ···················· | ···················· | ···················· | ···················· | ···················· |
| U | 6052 hCoV-19/Saudi Arabia/KAUST-MAKKAH201/2020\|EPI_ISL_ | ···················· | ···················· | ···················· | ···················· | ···················· | ···················· |
| U | 6053 hCoV-19/Saudi Arabia/KAUST-MAKKAH206/2020\|EPI_ISL_ | ···················· | ···················· | ···················· | ···················· | ···················· | ···················· |
| U | 6054 hCoV-19/Saudi Arabia/KAUST-MAKKAH222/2020\|EPI_ISL_ | ···················· | ···················· | ···················· | ···················· | ···················· | ···················· |
| U | 6055 hCoV-19/Saudi Arabia/KAUST-MAKKAH235/2020\|EPI_ISL_ | ···················· | ···················· | ···················· | ···················· | ···················· | ···················· |
| U | 6056 hCoV-19/Saudi Arabia/KAUST-MAKKAH26/2020\|EPI_ISL_6 | ···················· | ···················· | ···················· | ···················· | ···················· | ···················· |
| U | 6057 hCoV-19/Saudi Arabia/KAUST-MAKKAH729/2020\|EPI_ISL_ | ···················· | ·A·················· | ···················· | ···················· | ···················· | ···················· |
| U | 6058 hCoV-19/Saudi Arabia/KAUST-MAKKAH775/2020\|EPI_ISL_ | ···················· | ···················· | ···················· | ···················· | ···················· | ···················· |
| U | 6059 hCoV-19/Saudi Arabia/KAUST-MAKKAH798/2020\|EPI_ISL_ | ···················· | ···················· | ···················· | ···················· | ···················· | ···················· |
| U | 6060 hCoV-19/Saudi Arabia/KAUST-MAKKAH819/2020\|EPI_ISL_ | ···················· | ···················· | ···················· | ···················· | ···················· | ···················· |
| U | 6061 hCoV-19/Saudi Arabia/KAUST-MAKKAH825/2020\|EPI_ISL_ | ···················· | ···················· | ···················· | ···················· | ···················· | ···················· |
| U | 6062 hCoV-19/Saudi Arabia/KAUST-QATIF1344/2020\|EPI_ISL_ | ···················· | ·A·················· | ···················· | ···················· | ···················· | ···A················ |
| U | 6063 hCoV-19/Saudi Arabia/KAUST-QATIF1352/2020\|EPI_ISL_ | ···················· | ·A·················· | ···················· | ···················· | ···················· | ···A················ |
| U | 6064 hCoV-19/Saudi Arabia/KAUST-QATIF1353/2020\|EPI_ISL_ | ···················· | ·A·················· | ···················· | ···················· | ···················· | ···A················ |
| U | 6065 hCoV-19/Saudi Arabia/KAUST-QATIF1443/2020\|EPI_ISL_ | ···················· | ···················· | ···················· | ···················· | ···················· | ···A················ |
| U | 6066 hCoV-19/Saudi Arabia/KAUST-QATIF1448/2020\|EPI_ISL_ | ···················· | ···················· | ···················· | ···················· | ···················· | ···A················ |
| U | 6067 hCoV-19/Saudi Arabia/KAUST-QATIF1452/2020\|EPI_ISL_ | ···················· | ·A·················· | ···················· | ···················· | ···················· | ···A················ |
| U | 6068 hCoV-19/Saudi Arabia/KAUST-QATIF1457/2020\|EPI_ISL_ | ···················· | ···················· | ···················· | ···················· | ···················· | ···················· |
| U | 6069 hCoV-19/Saudi Arabia/KAUST-QATIF1468/2020\|EPI_ISL_ | ···················· | ···················· | ···················· | ···················· | ···················· | ···················· |
| U | 6070 hCoV-19/England/LOND-12517F7/2020\|EPI_ISL_679440\|2 | ···················· | ···················· | ···················· | ···················· | ···················· | ···················· |
| U | 6071 hCoV-19/England/LOND-1251803/2020\|EPI_ISL_679441\|2 | ···················· | ···················· | ···················· | ···················· | ···················· | ···················· |
| U | 6072 hCoV-19/England/LOND-12635AD/2020\|EPI_ISL_679450\|2 | ···················· | ···················· | ···················· | ···················· | ···················· | ···················· |
| U | 6073 hCoV-19/England/LOND-12635CB/2020\|EPI_ISL_679452\|2 | ···················· | ···················· | ···················· | ···················· | ···················· | ···················· |
| U | 6074 hCoV-19/England/LOND-12635E9/2020\|EPI_ISL_679454\|2 | ···················· | ···················· | ···················· | ···················· | ···················· | ···················· |
| U | 6075 hCoV-19/England/LOND-1263622/2020\|EPI_ISL_679458\|2 | ···················· | ···················· | ···················· | ···················· | ···················· | ···················· |
| U | 6076 hCoV-19/England/LOND-126368C/2020\|EPI_ISL_679463\|2 | ···················· | ···················· | ···················· | ···················· | ···················· | ···················· |
| U | 6077 hCoV-19/England/LOND-12636C8/2020\|EPI_ISL_679466\|2 | ···················· | ···················· | ···················· | ···················· | ···················· | ···················· |
| U | 6078 hCoV-19/England/LOND-126372F/2020\|EPI_ISL_679471\|2 | ···················· | ···················· | ···················· | ···················· | ···················· | ···················· |
| U | 6079 hCoV-19/England/LOND-126374D/2020\|EPI_ISL_679473\|2 | ···················· | ···················· | ···················· | ···················· | ···················· | ···················· |
| U | 6080 hCoV-19/England/LOND-12637A7/2020\|EPI_ISL_679476\|2 | ···················· | ···················· | ···················· | ···················· | ···················· | ···················· |
| U | 6081 hCoV-19/Northern Ireland/NIRE-234757/2020\|EPI_ISL_ | ···················· | ···················· | ···················· | ···················· | ···················· | ···················· |
| U | 6083 hCoV-19/Northern Ireland/NIRE-2347EE/2020\|EPI_ISL_ | ···················· | ···················· | ···················· | ···················· | ···················· | ···················· |
| U | 6084 hCoV-19/Northern Ireland/NIRE-2347FD/2020\|EPI_ISL_ | ···················· | ···················· | ···················· | ···················· | ···················· | ···················· |
| U | 6085 hCoV-19/Northern Ireland/NIRE-234818/2020\|EPI_ISL_ | ···················· | ···················· | ···················· | ···················· | ···················· | ···················· |
| U | 6086 hCoV-19/Northern Ireland/NIRE-234827/2020\|EPI_ISL_ | ···················· | ···················· | ···················· | ···················· | ···················· | ···················· |
| U | 6087 hCoV-19/Northern Ireland/NIRE-234845/2020\|EPI_ISL_ | ···················· | ·A·················· | ···················· | ···················· | ···················· | ···················· |
| U | 6088 hCoV-19/Scotland/CVR5715/2020\|EPI_ISL_680445\|2020- | ···················· | ···················· | ···················· | ···················· | ···················· | ···················· |
| U | 6089 hCoV-19/Scotland/CVR5727/2020\|EPI_ISL_680446\|2020- | ···················· | ···················· | ···················· | ···················· | ···················· | ···················· |
| U | 6090 hCoV-19/Scotland/CVR5798/2020\|EPI_ISL_680448\|2020- | ···················· | ···················· | ···················· | ···················· | ···················· | ···················· |
| U | 6091 hCoV-19/Scotland/CVR5870/2020\|EPI_ISL_680449\|2020- | ···················· | ···················· | ···················· | ···················· | ···················· | ···················· |
| U | 6092 hCoV-19/Scotland/CVR6008/2020\|EPI_ISL_680453\|2020- | ···················· | ···················· | ···················· | ···················· | ···················· | ···················· |
| U | 6093 hCoV-19/Scotland/CVR6021/2020\|EPI_ISL_680458\|2020- | ···················· | ···················· | ···················· | ···················· | ···················· | ···················· |

| | | | | | | | |
|---|---|---|---|---|---|---|---|
| U | 6094 hCoV-19/Scotland/CVR6022/2020\|EPI_ISL_680459\|2020- | ···················· | ···················· | ···················· | ···················· | ···················· | ···················· |
| U | 6095 hCoV-19/Scotland/EDB10044/2020\|EPI_ISL_680461\|2020 | ···················· | ···················· | ···················· | ···················· | ···················· | ···················· |
| U | 6096 hCoV-19/Scotland/EDB10046/2020\|EPI_ISL_680462\|2020 | ···················· | ···················· | ···················· | ···················· | ···················· | ···················· |
| U | 6100 hCoV-19/Scotland/EDB10083/2020\|EPI_ISL_680466\|2020 | ···················· | ···················· | ···················· | ···················· | ···················· | ···················· |
| U | 6101 hCoV-19/Scotland/EDB10089/2020\|EPI_ISL_680467\|2020 | ···················· | ···················· | ···················· | ···················· | ···················· | ···················· |
| U | 6103 hCoV-19/Scotland/EDB10104/2020\|EPI_ISL_680469\|2020 | ···················· | ···················· | ···················· | ···················· | ···················· | ···················· |
| U | 6104 hCoV-19/Scotland/EDB10105/2020\|EPI_ISL_680470\|2020 | ···················· | ···················· | ···················· | ···················· | ···················· | ···················· |
| U | 6105 hCoV-19/Scotland/EDB10107/2020\|EPI_ISL_680471\|2020 | ···················· | ···················· | ···················· | ···················· | ···················· | ···················· |
| U | 6107 hCoV-19/Chile/MA-CADIUMAG-35/2020\|EPI_ISL_681700\|2 | ···················· | ···················· | ···················· | ···················· | ···················· | ···················· |
| U | 6108 hCoV-19/Chile/MA-CADIUMAG-36/2020\|EPI_ISL_681701\|2 | ···················· | ···················· | ···················· | ···················· | ···················· | ···················· |
| U | 6109 hCoV-19/Ecuador/ZZ-SARS-2/2020\|EPI_ISL_681703\|2020 | ···················· | ···················· | ···················· | ···················· | ···················· | ···················· |
| U | 6110 hCoV-19/Ireland/CE-UHL135/2020\|EPI_ISL_681708\|2020 | ···················· | ···················· | ···················· | ···················· | ···················· | ···················· |
| U | 6111 hCoV-19/Ireland/CE-UHL23/2020\|EPI_ISL_681711\|2020- | ···················· | ···················· | ···················· | ···················· | ···················· | ···················· |
| U | 6112 hCoV-19/Ireland/CE-UHL298/2020\|EPI_ISL_681714\|2020 | ···················· | ···················· | ···················· | ···················· | ···················· | ···················· |
| U | 6113 hCoV-19/Ireland/CE-UHL58/2020\|EPI_ISL_681716\|2020- | ···················· | ···················· | ···················· | ···················· | ···················· | ···················· |
| U | 6114 hCoV-19/Ireland/LK-UHL8/2020\|EPI_ISL_681760\|2020-0 | ···················· | ···················· | ···················· | ···················· | ···················· | ···················· |
| U | 6115 hCoV-19/Ireland/TA-UHL11/2020\|EPI_ISL_681764\|2020- | ···················· | ···················· | ···················· | ···················· | ···················· | ···················· |
| U | 6116 hCoV-19/Ireland/TA-UHL2/2020\|EPI_ISL_681767\|2020-0 | ···················· | ···················· | ···················· | ···················· | ···················· | ···················· |
| U | 6117 hCoV-19/Ireland/TA-UHL22/2020\|EPI_ISL_681768\|2020- | ···················· | ···················· | ···················· | ···················· | ···················· | ···················· |
| U | 6118 hCoV-19/Ireland/TA-UHL25/2020\|EPI_ISL_681769\|2020- | ···················· | ···················· | ···················· | ···················· | ···················· | ···················· |
| U | 6119 hCoV-19/Ireland/TA-UHL40/2020\|EPI_ISL_681773\|2020- | ···················· | ···················· | ···················· | ···················· | ···················· | ···················· |
| U | 6120 hCoV-19/USA/GA-CDC-7327/2020\|EPI_ISL_681776\|2020-1 | ···················· | ···················· | ···················· | ···················· | ···················· | ···················· |
| U | 6121 hCoV-19/USA/GA-CDC-7328/2020\|EPI_ISL_681777\|2020-1 | ···················· | ···················· | ···················· | ···················· | ···················· | ···················· |
| U | 6122 hCoV-19/USA/GA-CDC-7329/2020\|EPI_ISL_681778\|2020-1 | ···················· | ···················· | ···················· | ···················· | ···················· | ···················· |
| U | 6123 hCoV-19/USA/GA-CDC-7345/2020\|EPI_ISL_681779\|2020-1 | ···················· | ···················· | ···················· | ···················· | ···················· | ···················· |
| U | 6124 hCoV-19/USA/GA-CDC-7347/2020\|EPI_ISL_681780\|2020-1 | ···················· | ···················· | ···················· | ···················· | ···················· | ···················· |
| U | 6125 hCoV-19/USA/GA-CDC-7361/2020\|EPI_ISL_681781\|2020-1 | ···················· | ···················· | ···················· | ···················· | ···················· | ···················· |
| U | 6126 hCoV-19/USA/GA-CDC-7364/2020\|EPI_ISL_681782\|2020-1 | ···················· | ···················· | ···················· | ···················· | ···················· | ···················· |
| U | 6127 hCoV-19/USA/GA-CDC-7365/2020\|EPI_ISL_681783\|2020-1 | ···················· | ···················· | ···················· | ···················· | ···················· | ···················· |
| U | 6128 hCoV-19/USA/GA-CDC-7366/2020\|EPI_ISL_681784\|2020-1 | ···················· | ···················· | ···················· | ···················· | ···················· | ···················· |
| U | 6129 hCoV-19/USA/GA-CDC-7369/2020\|EPI_ISL_681785\|2020-1 | ···················· | ···················· | ···················· | ···················· | ···················· | ···················· |
| U | 6130 hCoV-19/USA/GA-CDC-7371/2020\|EPI_ISL_681786\|2020-1 | ···················· | ···················· | ···················· | ···················· | ···················· | ···················· |
| U | 6131 hCoV-19/USA/GA-CDC-7378/2020\|EPI_ISL_681787\|2020-1 | ···················· | ···················· | ···················· | ···················· | ···················· | ···················· |
| U | 6132 hCoV-19/USA/GA-CDC-7388/2020\|EPI_ISL_681788\|2020-1 | ···················· | ···················· | ···················· | ···················· | ···················· | ···················· |
| U | 6133 hCoV-19/USA/GA-CDC-7492/2020\|EPI_ISL_681789\|2020-1 | ···················· | ···················· | ···················· | ···················· | ···················· | ···················· |
| U | 6134 hCoV-19/USA/GA-CDC-7516/2020\|EPI_ISL_681790\|2020-1 | ···················· | ···················· | ···················· | ···················· | ···················· | ···················· |
| U | 6135 hCoV-19/USA/GA-CDC-7519/2020\|EPI_ISL_681791\|2020-1 | ···················· | ···················· | ···················· | ···················· | ···················· | ···················· |
| U | 6136 hCoV-19/USA/GA-CDC-7525/2020\|EPI_ISL_681792\|2020-1 | ···················· | ···················· | ···················· | ···················· | ···················· | ···················· |
| U | 6137 hCoV-19/USA/GA-CDC-7533/2020\|EPI_ISL_681793\|2020-1 | ···················· | ···················· | ···················· | ···················· | ···················· | ···················· |
| U | 6138 hCoV-19/USA/GA-CDC-7543/2020\|EPI_ISL_681794\|2020-1 | ···················· | ···················· | ···················· | ···················· | ···················· | ···················· |
| U | 6139 hCoV-19/USA/GA-CDC-7556/2020\|EPI_ISL_681795\|2020-1 | ···················· | ···················· | ···················· | ···················· | ···················· | ···················· |
| U | 6140 hCoV-19/USA/GA-CDC-7561/2020\|EPI_ISL_681796\|2020-1 | ···················· | ···················· | ···················· | ···················· | ···················· | ···················· |
| U | 6141 hCoV-19/USA/GA-CDC-7591/2020\|EPI_ISL_681797\|2020-1 | ···················· | ···················· | ···················· | ···················· | ···················· | ···················· |
| U | 6142 hCoV-19/USA/GA-CDC-7595/2020\|EPI_ISL_681798\|2020-1 | ···················· | ···················· | ···················· | ···················· | ···················· | ···················· |
| U | 6143 hCoV-19/USA/GA-CDC-7596/2020\|EPI_ISL_681799\|2020-1 | ···················· | ···················· | ···················· | ···················· | ···················· | ···················· |
| U | 6144 hCoV-19/USA/UT-UPHL-201221794/2020\|EPI_ISL_684003\| | ···················· | ·A·················· | ···················· | ···················· | ···················· | ···················· |
| U | 6145 hCoV-19/England/CAMC-B09F5A/2020\|EPI_ISL_673224\|20 | ···················· | ···················· | ···················· | ···················· | ···················· | ···················· |
| U | 6146 hCoV-19/England/CAMC-B094A6/2020\|EPI_ISL_673225\|20 | ···················· | ···················· | ···················· | ···················· | ···················· | ···················· |
| U | 6147 hCoV-19/England/CAMC-B09BDE/2020\|EPI_ISL_673227\|20 | ···················· | ···················· | ···················· | ···················· | ···················· | ···················· |
| U | 6148 hCoV-19/England/CAMC-B09D8D/2020\|EPI_ISL_673228\|20 | ···················· | ···················· | ···················· | ···················· | ···················· | ···················· |
| U | 6149 hCoV-19/England/CAMC-B09B65/2020\|EPI_ISL_673229\|20 | ···················· | ···················· | ···················· | ···················· | ···················· | ···················· |
| U | 6150 hCoV-19/England/CAMC-B09066/2020\|EPI_ISL_673233\|20 | ···················· | ···················· | ···················· | ···················· | ···················· | ···················· |
| U | 6151 hCoV-19/England/CAMC-B08E7C/2020\|EPI_ISL_673234\|20 | ···················· | ···················· | ···················· | ···················· | ···················· | ···················· |
| U | 6152 hCoV-19/England/CAMC-B99748/2020\|EPI_ISL_673237\|20 | ···················· | ···················· | ···················· | ···················· | ···················· | ···················· |
| U | 6153 hCoV-19/England/CAMC-B9AAD5/2020\|EPI_ISL_673239\|20 | ···················· | ···················· | ···················· | ···················· | ······T············· | ···················· |
| U | 6154 hCoV-19/England/CAMC-B9ADAF/2020\|EPI_ISL_673240\|20 | ···················· | ···················· | ···················· | ···················· | ···················· | ···················· |
| U | 6155 hCoV-19/England/CAMC-B995A8/2020\|EPI_ISL_673241\|20 | ···················· | ···················· | ···················· | ···················· | ···················· | ···················· |
| U | 6156 hCoV-19/England/CAMC-B9ABA5/2020\|EPI_ISL_673242\|20 | ···················· | ···················· | ···················· | ···················· | ···················· | ···················· |
| U | 6157 hCoV-19/England/CAMC-B9AC0C/2020\|EPI_ISL_673244\|20 | ···················· | ···················· | ···················· | ···················· | ···················· | ···················· |
| U | 6158 hCoV-19/England/CAMC-B9C5E1/2020\|EPI_ISL_673245\|20 | ···················· | ···················· | ···················· | ···················· | ······T············· | ···················· |
| U | 6159 hCoV-19/England/CAMC-B9A422/2020\|EPI_ISL_673246\|20 | ···················· | ···················· | ···················· | ···················· | ···················· | ···················· |
| U | 6160 hCoV-19/England/CAMC-B99B4C/2020\|EPI_ISL_673247\|20 | ···················· | ···················· | ···················· | ···················· | ···················· | ···················· |
| U | 6161 hCoV-19/England/CAMC-B9939F/2020\|EPI_ISL_673248\|20 | ···················· | ···················· | ···················· | ···················· | ···················· | ···················· |
| U | 6162 hCoV-19/England/CAMC-B99C3A/2020\|EPI_ISL_673249\|20 | ···················· | ······A············· | ···················· | ···················· | ···················· | ···················· |
| U | 6163 hCoV-19/England/CAMC-B9A8AE/2020\|EPI_ISL_673250\|20 | ···················· | ···················· | ···················· | ···················· | ···················· | ···················· |
| U | 6164 hCoV-19/England/CAMC-B99793/2020\|EPI_ISL_673251\|20 | ···················· | ···················· | ···················· | ···················· | ···················· | ···················· |
| U | 6165 hCoV-19/England/CAMC-B9C4F3/2020\|EPI_ISL_673252\|20 | ···················· | ···················· | ···················· | ···················· | ···················· | ···················· |
| U | 6166 hCoV-19/England/CAMC-B9C4C6/2020\|EPI_ISL_673253\|20 | ···················· | ···················· | ···················· | ···················· | ···················· | ···················· |
| U | 6167 hCoV-19/England/CAMC-B9A3E9/2020\|EPI_ISL_673255\|20 | ···················· | ···················· | ···················· | ···················· | ···················· | ···················· |
| U | 6168 hCoV-19/England/CAMC-B99DDD/2020\|EPI_ISL_673256\|20 | ···················· | ···················· | ···················· | ···················· | ···················· | ···················· |
| U | 6169 hCoV-19/England/CAMC-B99FD7/2020\|EPI_ISL_673258\|20 | ···················· | ·A·················· | ···················· | ···················· | ···················· | ···················· |
| U | 6170 hCoV-19/England/CAMC-B9AAC6/2020\|EPI_ISL_673259\|20 | ···················· | ···················· | ···················· | ···················· | ···················· | ···················· |
| U | 6171 hCoV-19/England/CAMC-B98E9F/2020\|EPI_ISL_673260\|20 | ···················· | ···················· | ···················· | ···················· | ···················· | ···················· |
| U | 6172 hCoV-19/England/CAMC-B98F32/2020\|EPI_ISL_673261\|20 | ···················· | ···················· | ···················· | ···················· | ···················· | ···················· |
| U | 6173 hCoV-19/England/CAMC-B9AA8A/2020\|EPI_ISL_673262\|20 | ···················· | ···················· | ···················· | ···················· | ···················· | ···················· |
| U | 6174 hCoV-19/England/CAMC-B9A99C/2020\|EPI_ISL_673266\|20 | ···················· | ···················· | ···················· | ···················· | ···················· | ···················· |
| U | 6175 hCoV-19/England/CAMC-B9A3F8/2020\|EPI_ISL_673267\|20 | ···················· | ···················· | ···················· | ···················· | ···················· | ···················· |
| U | 6176 hCoV-19/England/QEUH-B7F54E/2020\|EPI_ISL_674501\|20 | ···················· | ···················· | ···················· | ···················· | ···················· | ···················· |
| U | 6177 hCoV-19/England/MILK-B8EBFF/2020\|EPI_ISL_674503\|20 | ···················· | ···················· | ···················· | ···················· | ···················· | ···················· |
| U | 6178 hCoV-19/England/MILK-B8EED8/2020\|EPI_ISL_674505\|20 | ···················· | ···················· | ···················· | ···················· | ···················· | ···················· |

| | | | | | | | | |
|---|---|---|---|---|---|---|---|---|
| U | 6179 | hCoV-19/England/MILK-B8E3D9/2020\|EPI_ISL_674507\|20 | ···················· | ·A·················· | ···················· | ···················· | ···················· | ···A················ |
| U | 6180 | hCoV-19/England/MILK-B8ED9F/2020\|EPI_ISL_674508\|20 | ···················· | ···················· | ···················· | ···················· | ···················· | ···················· |
| U | 6181 | hCoV-19/England/MILK-B8F7FA/2020\|EPI_ISL_674509\|20 | ···················· | ···················· | ···················· | ···················· | ···················· | ···················· |
| U | 6182 | hCoV-19/England/MILK-B8ED53/2020\|EPI_ISL_674510\|20 | ···················· | ···················· | ···················· | ···················· | ···················· | ···················· |
| U | 6183 | hCoV-19/England/MILK-B8F8CA/2020\|EPI_ISL_674555\|20 | ···················· | ···················· | ···················· | ···················· | ···················· | ···················· |
| U | 6184 | hCoV-19/England/MILK-B8F824/2020\|EPI_ISL_674556\|20 | ···················· | ···················· | ···················· | ···················· | ···················· | ···················· |
| U | 6185 | hCoV-19/England/QEUH-B808F6/2020\|EPI_ISL_674558\|20 | ···················· | ···················· | ···················· | ···················· | ···················· | ···················· |
| U | 6186 | hCoV-19/England/QEUH-B7F57B/2020\|EPI_ISL_674563\|20 | ···················· | ···················· | ···················· | ···················· | ···················· | ···················· |
| U | 6187 | hCoV-19/England/MILK-B8EAC5/2020\|EPI_ISL_674564\|20 | ···················· | ···················· | ···················· | ···················· | ···················· | ···················· |
| U | 6188 | hCoV-19/England/MILK-B8F5C3/2020\|EPI_ISL_674566\|20 | ···················· | ···················· | ···················· | ···················· | ···················· | ···················· |
| U | 6189 | hCoV-19/England/MILK-B8EE8D/2020\|EPI_ISL_674567\|20 | ···················· | ···················· | ···················· | ···················· | ···················· | ···················· |
| U | 6190 | hCoV-19/England/MILK-B8DFD6/2020\|EPI_ISL_674568\|20 | ···················· | ···················· | ···················· | ···················· | ···················· | ···················· |
| U | 6191 | hCoV-19/England/QEUH-B8095D/2020\|EPI_ISL_674569\|20 | ···················· | ···················· | ···················· | ···················· | ···················· | ···················· |
| U | 6192 | hCoV-19/England/MILK-B8E4F4/2020\|EPI_ISL_674570\|20 | ···················· | ···················· | ···················· | ···················· | ···················· | ···················· |
| U | 6193 | hCoV-19/England/MILK-B8EC29/2020\|EPI_ISL_674572\|20 | ···················· | ···················· | ···················· | ···················· | ···················· | ···················· |
| U | 6194 | hCoV-19/England/MILK-B8F781/2020\|EPI_ISL_674573\|20 | ···················· | ···················· | ···················· | ···················· | ···················· | ···················· |
| U | 6195 | hCoV-19/England/MILK-B8F596/2020\|EPI_ISL_674574\|20 | ···················· | ···················· | ···················· | ···················· | ···················· | ···················· |
| U | 6196 | hCoV-19/England/MILK-B8ED26/2020\|EPI_ISL_674575\|20 | ···················· | ···················· | ···················· | ···················· | ···················· | ···················· |
| U | 6197 | hCoV-19/England/MILK-B8F61B/2020\|EPI_ISL_674577\|20 | ···················· | ···················· | ···················· | ···················· | ···················· | ···················· |
| U | 6198 | hCoV-19/England/MILK-B8EC47/2020\|EPI_ISL_674578\|20 | ···················· | ···················· | ···················· | ···················· | ···················· | ···················· |
| U | 6199 | hCoV-19/England/MILK-B8DFB8/2020\|EPI_ISL_674580\|20 | ···················· | ···················· | ···················· | ···················· | ···················· | ···················· |
| U | 6200 | hCoV-19/England/MILK-B8F587/2020\|EPI_ISL_674581\|20 | ···················· | ···················· | ···················· | ···················· | ···················· | ···················· |
| U | 6201 | hCoV-19/England/MILK-B8DFA9/2020\|EPI_ISL_674582\|20 | ···················· | ···················· | ···················· | ···················· | ···················· | ···A················ |
| U | 6202 | hCoV-19/England/QEUH-B7F71B/2020\|EPI_ISL_674585\|20 | ···················· | ·A·················· | ···················· | ···················· | ···················· | ···················· |
| U | 6203 | hCoV-19/England/MILK-B8F709/2020\|EPI_ISL_674586\|20 | ···················· | ···················· | ···················· | ···················· | ···················· | ···················· |
| U | 6204 | hCoV-19/England/MILK-B8EEAB/2020\|EPI_ISL_674587\|20 | ···················· | ···················· | ···················· | ···················· | ···················· | ···················· |
| U | 6205 | hCoV-19/England/QEUH-B7D6D4/2020\|EPI_ISL_674589\|20 | ···················· | ···················· | ···················· | ···················· | ···················· | ···················· |
| U | 6206 | hCoV-19/England/QEUH-B7CF34/2020\|EPI_ISL_674590\|20 | ···················· | ···················· | ···················· | ···················· | ···················· | ···················· |
| U | 6207 | hCoV-19/England/MILK-B8F44E/2020\|EPI_ISL_674591\|20 | ···················· | ···················· | ···················· | ···················· | ···················· | ···················· |
| U | 6208 | hCoV-19/England/MILK-B8EAA7/2020\|EPI_ISL_674593\|20 | ···················· | ···················· | ···················· | ···················· | ···················· | ···················· |
| U | 6209 | hCoV-19/England/MILK-B8DDBE/2020\|EPI_ISL_674594\|20 | ···················· | ···················· | ···················· | ···················· | ···················· | ···················· |
| U | 6210 | hCoV-19/England/QEUH-B7D6B6/2020\|EPI_ISL_674595\|20 | ···················· | ···················· | ···················· | ···················· | ···················· | ···················· |
| U | 6211 | hCoV-19/England/MILK-B8E807/2020\|EPI_ISL_674596\|20 | ···················· | ···················· | ···················· | ···················· | ···················· | ···················· |
| U | 6212 | hCoV-19/England/MILK-B8DF9A/2020\|EPI_ISL_674597\|20 | ···················· | ···················· | ···················· | ···················· | ···················· | ···················· |
| U | 6213 | hCoV-19/England/MILK-B8F48A/2020\|EPI_ISL_674598\|20 | ···················· | ·A·······A·········· | ···················· | ···················· | ···················· | ···················· |
| U | 6214 | hCoV-19/England/ALDP-B84B80/2020\|EPI_ISL_675800\|20 | ···················· | ···················· | ···················· | ···················· | ···················· | ···················· |
| U | 6215 | hCoV-19/England/ALDP-B84FFD/2020\|EPI_ISL_675802\|20 | ···················· | ···················· | ···················· | ···················· | ···················· | ···················· |
| U | 6216 | hCoV-19/England/ALDP-B848F2/2020\|EPI_ISL_675803\|20 | ···················· | ···················· | ···················· | ···················· | ···················· | ···················· |
| U | 6217 | hCoV-19/England/MILK-B889DD/2020\|EPI_ISL_675804\|20 | ···················· | ···················· | ···················· | ···················· | ···················· | ···················· |
| U | 6218 | hCoV-19/England/MILK-B88C98/2020\|EPI_ISL_675805\|20 | ···················· | ···················· | ···················· | ···················· | ···················· | ···················· |
| U | 6219 | hCoV-19/England/ALDP-B84810/2020\|EPI_ISL_675806\|20 | ···················· | ···················· | ···················· | ···················· | ······T············· | ···················· |
| U | 6220 | hCoV-19/England/MILK-B88982/2020\|EPI_ISL_675807\|20 | ···················· | ···················· | ···················· | ···················· | ···················· | ···················· |
| U | 6221 | hCoV-19/England/MILK-B8C8DC/2020\|EPI_ISL_675808\|20 | ···················· | ···················· | ···················· | ···················· | ···················· | ···················· |
| U | 6222 | hCoV-19/England/MILK-B8C739/2020\|EPI_ISL_675809\|20 | ···················· | ···················· | ···················· | ···················· | ···················· | ···················· |
| U | 6223 | hCoV-19/England/ALDP-B84B62/2020\|EPI_ISL_675810\|20 | ···················· | ···················· | ···················· | ···················· | ······T············· | ···················· |
| U | 6224 | hCoV-19/England/MILK-B899EB/2020\|EPI_ISL_675811\|20 | ···················· | ···················· | ···················· | ···················· | ···················· | ···················· |
| U | 6225 | hCoV-19/England/ALDP-B84D5D/2020\|EPI_ISL_675812\|20 | ···················· | ···················· | ···················· | ···················· | ···················· | ···················· |
| U | 6226 | hCoV-19/England/MILK-B8CA21/2020\|EPI_ISL_675814\|20 | ···················· | ···················· | ···················· | ···················· | ···················· | ···················· |
| U | 6227 | hCoV-19/England/MILK-B8890A/2020\|EPI_ISL_675815\|20 | ···················· | ···················· | ···················· | ···················· | ···················· | ···················· |
| U | 6228 | hCoV-19/England/MILK-B88BC8/2020\|EPI_ISL_675816\|20 | ···················· | ···················· | ···················· | ···················· | ···················· | ···················· |
| U | 6229 | hCoV-19/England/ALDP-B84D99/2020\|EPI_ISL_675817\|20 | ···················· | ···················· | ···················· | ···················· | ···················· | ···················· |
| U | 6230 | hCoV-19/England/ALDP-B84CF6/2020\|EPI_ISL_675818\|20 | ···················· | ···················· | ···················· | ···················· | ···················· | ···················· |
| U | 6231 | hCoV-19/England/MILK-B8973C/2020\|EPI_ISL_675819\|20 | ···················· | ···················· | ···················· | ···················· | ···················· | ···················· |
| U | 6232 | hCoV-19/England/ALDP-B84D7B/2020\|EPI_ISL_675820\|20 | ···················· | ···················· | ···················· | ···················· | ···················· | ···················· |
| U | 6233 | hCoV-19/England/MILK-B89787/2020\|EPI_ISL_675821\|20 | ···················· | ···················· | ···················· | ···················· | ···················· | ···················· |
| U | 6234 | hCoV-19/England/ALDP-B8490E/2020\|EPI_ISL_675823\|20 | ···················· | ···················· | ···················· | ···················· | ···················· | ···················· |
| U | 6235 | hCoV-19/England/MILK-B88964/2020\|EPI_ISL_675824\|20 | ···················· | ·A·················· | ···················· | ···················· | ···················· | ···················· |
| U | 6236 | hCoV-19/England/ALDP-B84E3C/2020\|EPI_ISL_675825\|20 | ···················· | ·A·················· | ···················· | ···················· | ···················· | ···A················ |
| U | 6237 | hCoV-19/England/ALDP-B84E69/2020\|EPI_ISL_675826\|20 | ···················· | ···················· | ···················· | ···················· | ···················· | ···················· |
| U | 6238 | hCoV-19/England/MILK-B88B5F/2020\|EPI_ISL_675828\|20 | ···················· | ···················· | ···················· | ···················· | ···················· | ···················· |
| U | 6239 | hCoV-19/England/ALDP-B84BBD/2020\|EPI_ISL_675829\|20 | ···················· | ···················· | ···················· | ···················· | ···················· | ···················· |
| U | 6240 | hCoV-19/England/ALDP-B859EF/2020\|EPI_ISL_675833\|20 | ···················· | ···················· | ···················· | ···················· | ···················· | ···················· |
| U | 6241 | hCoV-19/England/MILK-B8C6E1/2020\|EPI_ISL_675834\|20 | ···················· | ···················· | ···················· | ···················· | ···················· | ···················· |
| U | 6242 | hCoV-19/England/MILK-B8CB00/2020\|EPI_ISL_675835\|20 | ···················· | ···················· | ···················· | ···················· | ···················· | ···················· |
| U | 6243 | hCoV-19/England/MILK-B8970F/2020\|EPI_ISL_675837\|20 | ···················· | ·A·················· | ···················· | ···················· | ···················· | ···A················ |
| U | 6244 | hCoV-19/England/MILK-B88C10/2020\|EPI_ISL_675838\|20 | ···················· | ···················· | ···················· | ···················· | ···················· | ···················· |
| U | 6245 | hCoV-19/England/ALDP-B850EA/2020\|EPI_ISL_675839\|20 | ···················· | ···················· | ···················· | ···················· | ···················· | ···················· |
| U | 6246 | hCoV-19/England/MILK-B89699/2020\|EPI_ISL_675840\|20 | ···················· | ···················· | ···················· | ···················· | ···················· | ···················· |
| U | 6247 | hCoV-19/England/MILK-B8CA7C/2020\|EPI_ISL_675886\|20 | ···················· | ···················· | ···················· | ···················· | ···················· | ···················· |
| U | 6248 | hCoV-19/England/MILK-B88B40/2020\|EPI_ISL_675888\|20 | ···················· | ···················· | ···················· | ···················· | ···················· | ···················· |
| U | 6249 | hCoV-19/England/MILK-B89796/2020\|EPI_ISL_675891\|20 | ···················· | ···················· | ···················· | ···················· | ···················· | ···················· |
| U | 6250 | hCoV-19/England/ALDP-B84DB7/2020\|EPI_ISL_675892\|20 | ···················· | ···················· | ···················· | ···················· | ···················· | ···················· |
| U | 6251 | hCoV-19/England/MILK-B889FB/2020\|EPI_ISL_675893\|20 | ···················· | ···················· | ···················· | ···················· | ···················· | ···················· |
| U | 6252 | hCoV-19/England/ALDP-B84E78/2020\|EPI_ISL_675894\|20 | ···················· | ···················· | ···················· | ···················· | ···················· | ···················· |
| U | 6253 | hCoV-19/England/ALDP-B8484D/2020\|EPI_ISL_675897\|20 | ···················· | ·A·················· | ···················· | ···················· | ···················· | ···A················ |
| U | 6254 | hCoV-19/England/MILK-B899FA/2020\|EPI_ISL_675898\|20 | ···················· | ···················· | ···················· | ···················· | ···················· | ···················· |
| U | 6255 | hCoV-19/England/MILK-B89769/2020\|EPI_ISL_675899\|20 | ···················· | ···················· | ···················· | ···················· | ···················· | ···················· |
| U | 6256 | hCoV-19/Saudi Arabia/KAUST-MAKKAH1413/2020\|EPI_ISL | ···················· | ···················· | ···················· | ···················· | ···················· | ···················· |
| U | 6257 | hCoV-19/Saudi Arabia/KAUST-MAKKAH176/2020\|EPI_ISL_ | ···················· | ···················· | ···················· | ···················· | ···················· | ···················· |
| U | 6258 | hCoV-19/Saudi Arabia/KAUST-MAKKAH191/2020\|EPI_ISL_ | ···················· | ···················· | ···················· | ···················· | ···················· | ···················· |

| | | | | | | | | | |
|---|---|---|---|---|---|---|---|---|---|
| U | 6259 | hCoV-19/Saudi Arabia/KAUST-MAKKAH221/2020\|EPI_ISL_ | ···················· | ···················· | ····················· | ···················· | ···················· | ···················· | |
| U | 6260 | hCoV-19/Saudi Arabia/KAUST-MAKKAH118/2020\|EPI_ISL_ | ···················· | ···················· | ····················· | ···················· | ···················· | ···················· | |
| U | 6262 | hCoV-19/Saudi Arabia/KAUST-MAKKAH104/2020\|EPI_ISL_ | ···················· | ···················· | ····················· | ···················· | ···················· | ···················· | |
| U | 6263 | hCoV-19/Saudi Arabia/KAUST-MAKKAH106/2020\|EPI_ISL_ | ···················· | ···················· | ····················· | ···················· | ···················· | ···················· | |
| U | 6264 | hCoV-19/Saudi Arabia/KAUST-MAKKAH11/2020\|EPI_ISL_6 | ···················· | ···················· | ····················· | ···················· | ···················· | ···················· | |
| U | 6265 | hCoV-19/Saudi Arabia/KAUST-MAKKAH112/2020\|EPI_ISL_ | ···················· | ···················· | ····················· | ···················· | ···················· | ···················· | |
| U | 6266 | hCoV-19/Saudi Arabia/KAUST-MAKKAH117/2020\|EPI_ISL_ | ···················· | ···················· | ····················· | ···················· | ···················· | ···················· | |
| U | 6267 | hCoV-19/Saudi Arabia/KAUST-MAKKAH1388/2020\|EPI_ISL | ···················· | ·A·················· | ····················· | ···················· | ···················· | ···A················ | |
| U | 6268 | hCoV-19/Saudi Arabia/KAUST-MAKKAH1392/2020\|EPI_ISL | ···················· | ·A·················· | ····················· | ···················· | ···················· | ···A················ | |
| U | 6269 | hCoV-19/Saudi Arabia/KAUST-MAKKAH1395/2020\|EPI_ISL | ···················· | ·A·················· | ····················· | ···················· | ···················· | ···A················ | |
| U | 6270 | hCoV-19/Saudi Arabia/KAUST-MAKKAH1399/2020\|EPI_ISL | ···················· | ·A·················· | ····················· | ···················· | ···················· | ···A················ | |
| U | 6271 | hCoV-19/Saudi Arabia/KAUST-MAKKAH1401/2020\|EPI_ISL | ···················· | ·A·················· | ····················· | ···················· | ···················· | ···A················ | |
| U | 6272 | hCoV-19/Saudi Arabia/KAUST-MAKKAH1404/2020\|EPI_ISL | ···················· | ···················· | ····················· | ···················· | ···················· | ···················· | |
| U | 6273 | hCoV-19/Saudi Arabia/KAUST-MAKKAH1406/2020\|EPI_ISL | ···················· | ·A·················· | ····················· | ···················· | ···················· | ···A················ | |
| U | 6274 | hCoV-19/Saudi Arabia/KAUST-MAKKAH1408/2020\|EPI_ISL | ···················· | ·A·················· | ····················· | ···················· | ···················· | ···A················ | |
| U | 6275 | hCoV-19/Saudi Arabia/KAUST-MAKKAH1410/2020\|EPI_ISL | ···················· | ···················· | ····················· | ···················· | ···················· | ···················· | |
| U | 6276 | hCoV-19/Saudi Arabia/KAUST-MAKKAH1415/2020\|EPI_ISL | ···················· | ···················· | ····················· | ···················· | ···················· | ···················· | |
| U | 6277 | hCoV-19/Saudi Arabia/KAUST-MAKKAH1421/2020\|EPI_ISL | ···················· | ···················· | ····················· | ···················· | ···················· | ···················· | |
| U | 6278 | hCoV-19/Saudi Arabia/KAUST-MAKKAH1426/2020\|EPI_ISL | ···················· | ···················· | ····················· | ···················· | ···················· | ···················· | |
| U | 6279 | hCoV-19/Saudi Arabia/KAUST-MAKKAH1427/2020\|EPI_ISL | ···················· | ···················· | ····················· | ···················· | ···················· | ···················· | |
| U | 6280 | hCoV-19/Saudi Arabia/KAUST-MAKKAH150/2020\|EPI_ISL_ | ···················· | ···················· | ····················· | ···················· | ···················· | ···················· | |
| U | 6281 | hCoV-19/Saudi Arabia/KAUST-MAKKAH1503/2020\|EPI_ISL | ···················· | ···················· | ····················· | ···················· | ···················· | ···················· | |
| U | 6282 | hCoV-19/Saudi Arabia/KAUST-MAKKAH1504/2020\|EPI_ISL | ···················· | ···················· | ····················· | ···················· | ···················· | ···················· | |
| U | 6283 | hCoV-19/Saudi Arabia/KAUST-MAKKAH1515/2020\|EPI_ISL | ···················· | ···················· | ····················· | ···················· | ···················· | ···················· | |
| U | 6284 | hCoV-19/Saudi Arabia/KAUST-MAKKAH1517/2020\|EPI_ISL | ···················· | ···················· | ····················· | ···················· | ···················· | ···················· | |
| U | 6285 | hCoV-19/Saudi Arabia/KAUST-MAKKAH1520/2020\|EPI_ISL | ···················· | ···················· | ····················· | ···················· | ···················· | ···················· | |
| U | 6286 | hCoV-19/Saudi Arabia/KAUST-MAKKAH1525/2020\|EPI_ISL | ···················· | ···················· | ····················· | ···················· | ···················· | ···················· | |
| U | 6287 | hCoV-19/Saudi Arabia/KAUST-MAKKAH1527/2020\|EPI_ISL | ···················· | ···················· | ····················· | ···················· | ···················· | ···················· | |
| U | 6288 | hCoV-19/Saudi Arabia/KAUST-MAKKAH1536/2020\|EPI_ISL | ···················· | ·A·················· | ····················· | ···················· | ···················· | ···A················ | |
| U | 6289 | hCoV-19/Saudi Arabia/KAUST-MAKKAH1537/2020\|EPI_ISL | ···················· | ·A·················· | ····················· | ···················· | ···················· | ···················· | |
| U | 6290 | hCoV-19/Saudi Arabia/KAUST-MAKKAH1539/2020\|EPI_ISL | ···················· | ···················· | ····················· | ···················· | ···················· | ···················· | |
| U | 6291 | hCoV-19/Saudi Arabia/KAUST-MAKKAH1542/2020\|EPI_ISL | ···················· | ···················· | ····················· | ···················· | ···················· | ···················· | |
| U | 6292 | hCoV-19/Saudi Arabia/KAUST-MAKKAH1549/2020\|EPI_ISL | ···················· | ···················· | ····················· | ···················· | ···················· | ···················· | |
| U | 6293 | hCoV-19/Saudi Arabia/KAUST-MAKKAH1554/2020\|EPI_ISL | ···················· | ···················· | ····················· | ···················· | ···················· | ···················· | |
| U | 6294 | hCoV-19/Saudi Arabia/KAUST-MAKKAH1558/2020\|EPI_ISL | ···················· | ···················· | ····················· | ···················· | ···················· | ···················· | |
| U | 6295 | hCoV-19/Saudi Arabia/KAUST-MAKKAH1559/2020\|EPI_ISL | ···················· | ···················· | ····················· | ···················· | ···················· | ···················· | |
| U | 6296 | hCoV-19/Saudi Arabia/KAUST-MAKKAH1560/2020\|EPI_ISL | ···················· | ···················· | ····················· | ···················· | ···················· | ···················· | |
| U | 6297 | hCoV-19/Saudi Arabia/KAUST-MAKKAH1562/2020\|EPI_ISL | ···················· | ···················· | ····················· | ···················· | ···················· | ···················· | |
| U | 6298 | hCoV-19/Saudi Arabia/KAUST-MAKKAH1566/2020\|EPI_ISL | ···················· | ···················· | ····················· | ···················· | ···················· | ···················· | |
| U | 6299 | hCoV-19/Saudi Arabia/KAUST-MAKKAH157/2020\|EPI_ISL_ | ···················· | ···················· | ····················· | ···················· | ···················· | ···················· | |
| U | 6300 | hCoV-19/Saudi Arabia/KAUST-MAKKAH158/2020\|EPI_ISL_ | ···················· | ···················· | ····················· | ···················· | ···················· | ···················· | |
| U | 6301 | hCoV-19/Saudi Arabia/KAUST-MAKKAH1588/2020\|EPI_ISL | ···················· | ···················· | ····················· | ···················· | ···················· | ···················· | |
| U | 6302 | hCoV-19/England/LIVE-DBD2E1/2020\|EPI_ISL_679391\|20 | ···················· | ···················· | ····················· | ···················· | ···················· | ···················· | |
| U | 6303 | hCoV-19/England/LOND-125150C/2020\|EPI_ISL_679407\|2 | ···················· | ·A·················· | ····················· | ···················· | ···················· | ···················· | |
| U | 6304 | hCoV-19/England/LOND-125152A/2020\|EPI_ISL_679408\|2 | ···················· | ···················· | ····················· | ···················· | ···················· | ···················· | |
| U | 6306 | hCoV-19/England/LOND-1251548/2020\|EPI_ISL_679410\|2 | ···················· | ···················· | ····················· | ···················· | ···················· | ···················· | |
| U | 6308 | hCoV-19/England/LOND-12515DF/2020\|EPI_ISL_679415\|2 | ···················· | ···················· | ····················· | ···················· | ···················· | ···················· | |
| U | 6309 | hCoV-19/England/LOND-1251618/2020\|EPI_ISL_679416\|2 | ···················· | ···················· | ····················· | ···················· | ······T············· | ···················· | |
| U | 6310 | hCoV-19/England/LOND-1251672/2020\|EPI_ISL_679421\|2 | ···················· | ···················· | ····················· | ···················· | ···················· | ···················· | |
| U | 6311 | hCoV-19/England/LOND-1251690/2020\|EPI_ISL_679423\|2 | ···················· | ···················· | ····················· | ···················· | ···················· | ···················· | |
| U | 6312 | hCoV-19/England/LOND-12516DC/2020\|EPI_ISL_679425\|2 | ···················· | ···················· | ····················· | ···················· | ······T············· | ···················· | |
| U | 6313 | hCoV-19/England/LOND-12516EB/2020\|EPI_ISL_679426\|2 | ···················· | ···················· | ····················· | ···················· | ···················· | ···················· | |
| U | 6314 | hCoV-19/England/LOND-12516FA/2020\|EPI_ISL_679427\|2 | ···················· | ···················· | ····················· | ···················· | ······T············· | ···················· | |
| U | 6315 | hCoV-19/England/LOND-1251724/2020\|EPI_ISL_679429\|2 | ···················· | ···················· | ····················· | ···················· | ······T············· | ···················· | |
| U | 6316 | hCoV-19/England/LOND-1251760/2020\|EPI_ISL_679432\|2 | ···················· | ···················· | ····················· | ···················· | ···················· | ···················· | |
| U | 6317 | hCoV-19/England/LOND-125178E/2020\|EPI_ISL_679434\|2 | ···················· | ···················· | ····················· | ···················· | ···················· | ···················· | |
| U | 6319 | hCoV-19/England/LOND-12517CA/2020\|EPI_ISL_679438\|2 | ···················· | ···················· | ····················· | ···················· | ···················· | ···················· | |
| U | 6320 | hCoV-19/England/LOND-125187C/2020\|EPI_ISL_679446\|2 | ···················· | ···················· | ····················· | ···················· | ···················· | ···················· | |
| U | 6321 | hCoV-19/England/LOND-126359E/2020\|EPI_ISL_679449\|2 | ···················· | ···················· | ····················· | ···················· | ···················· | ···················· | |
| U | 6322 | hCoV-19/Northern Ireland/NIRE-2341D1/2020\|EPI_ISL_ | ···················· | ···················· | ····················· | ···················· | ···················· | ···················· | |
| U | 6323 | hCoV-19/Northern Ireland/NIRE-2341E0/2020\|EPI_ISL_ | ···················· | ···················· | ····················· | ···················· | ···················· | ···················· | |
| U | 6324 | hCoV-19/Northern Ireland/NIRE-23421A/2020\|EPI_ISL_ | ···················· | ···················· | ····················· | ···················· | ···················· | ···················· | |
| U | 6325 | hCoV-19/Northern Ireland/NIRE-234229/2020\|EPI_ISL_ | ···················· | ···················· | ····················· | ···················· | ···················· | ···················· | |
| U | 6326 | hCoV-19/Northern Ireland/NIRE-234238/2020\|EPI_ISL_ | ···················· | ···················· | ····················· | ···················· | ···················· | ···················· | |
| U | 6327 | hCoV-19/Northern Ireland/NIRE-234274/2020\|EPI_ISL_ | ···················· | ···················· | ····················· | ···················· | ···················· | ···················· | |
| U | 6328 | hCoV-19/Northern Ireland/NIRE-234292/2020\|EPI_ISL_ | ···················· | ···················· | ····················· | ···················· | ···················· | ···················· | |
| U | 6329 | hCoV-19/Northern Ireland/NIRE-2342B0/2020\|EPI_ISL_ | ···················· | ···················· | ····················· | ···················· | ···················· | ···················· | |
| U | 6330 | hCoV-19/Northern Ireland/NIRE-2342CF/2020\|EPI_ISL_ | ···················· | ···················· | ····················· | ···················· | ···················· | ···················· | |
| U | 6331 | hCoV-19/Northern Ireland/NIRE-2342DE/2020\|EPI_ISL_ | ···················· | ···················· | ····················· | ···················· | ···················· | ···················· | |
| U | 6335 | hCoV-19/Northern Ireland/NIRE-234344/2020\|EPI_ISL_ | ···················· | ···················· | ····················· | ···················· | ···················· | ···················· | |
| U | 6337 | hCoV-19/Northern Ireland/NIRE-234371/2020\|EPI_ISL_ | ···················· | ···················· | ····················· | ···················· | ···················· | ···················· | |
| U | 6338 | hCoV-19/Northern Ireland/NIRE-234380/2020\|EPI_ISL_ | ···················· | ···················· | ····················· | ···················· | ···················· | ···················· | |
| U | 6340 | hCoV-19/Northern Ireland/NIRE-2343BD/2020\|EPI_ISL_ | ···················· | ···················· | ····················· | ···················· | ···················· | ···················· | |
| U | 6341 | hCoV-19/Northern Ireland/NIRE-2343CC/2020\|EPI_ISL_ | ··················T· | ···················· | ····················· | ···················· | ···················· | ···················· | 1 |
| U | 6342 | hCoV-19/Northern Ireland/NIRE-2343DB/2020\|EPI_ISL_ | ···················· | ···················· | ····················· | ···················· | ···················· | ···················· | |
| U | 6344 | hCoV-19/Northern Ireland/NIRE-2343F9/2020\|EPI_ISL_ | ···················· | ···················· | ····················· | ···················· | ···················· | ···················· | |
| U | 6345 | hCoV-19/Northern Ireland/NIRE-234405/2020\|EPI_ISL_ | ···················· | ···················· | ····················· | ···················· | ···················· | ···················· | |
| U | 6347 | hCoV-19/Northern Ireland/NIRE-234432/2020\|EPI_ISL_ | ···················· | ···················· | ····················· | ···················· | ···················· | ···················· | |
| U | 6348 | hCoV-19/Northern Ireland/NIRE-234441/2020\|EPI_ISL_ | ···················· | ···················· | ····················· | ···················· | ···················· | ···················· | |
| U | 6349 | hCoV-19/Northern Ireland/NIRE-234450/2020\|EPI_ISL_ | ···················· | ···················· | ····················· | ···················· | ···················· | ···················· | |

| | | | | | | | | | |
|---|---|---|---|---|---|---|---|---|---|
| U | 6350 | hCoV-19/Northern Ireland/NIRE-23446F/2020\|EPI_ISL_ | .................... | .................... | .................... | .................... | .................... | .................... | |
| U | 6351 | hCoV-19/Northern Ireland/NIRE-23447E/2020\|EPI_ISL_ | .................... | .................... | .................... | .................... | .................... | .................... | |
| U | 6352 | hCoV-19/Northern Ireland/NIRE-23448D/2020\|EPI_ISL_ | .................... | .................... | .................... | .................... | .................... | .................... | |
| U | 6353 | hCoV-19/Northern Ireland/NIRE-23449C/2020\|EPI_ISL_ | .................... | .................... | .................... | .................... | .................... | .................... | |
| U | 6354 | hCoV-19/Northern Ireland/NIRE-2344AB/2020\|EPI_ISL_ | .................... | .................... | .................... | .................... | .................... | .................... | |
| U | 6355 | hCoV-19/Northern Ireland/NIRE-2344BA/2020\|EPI_ISL_ | .................... | .................... | .................... | .................... | .................... | .................... | |
| U | 6356 | hCoV-19/Northern Ireland/NIRE-2344C9/2020\|EPI_ISL_ | .................... | .................... | .................... | .................... | .................... | .................... | |
| U | 6358 | hCoV-19/Northern Ireland/NIRE-2344E7/2020\|EPI_ISL_ | .................... | .................... | .................... | .................... | .................... | .................... | |
| U | 6360 | hCoV-19/Northern Ireland/NIRE-234502/2020\|EPI_ISL_ | .................... | .................... | .................... | .................... | .................... | .................... | |
| U | 6361 | hCoV-19/Northern Ireland/NIRE-234520/2020\|EPI_ISL_ | .................... | .................... | .................... | .................... | .................... | .................... | |
| U | 6362 | hCoV-19/Northern Ireland/NIRE-23453F/2020\|EPI_ISL_ | .................... | .................... | .................... | .................... | .................... | .................... | |
| U | 6363 | hCoV-19/Northern Ireland/NIRE-23454E/2020\|EPI_ISL_ | .................... | .................... | .................... | .................... | .................... | .................... | |
| U | 6364 | hCoV-19/Northern Ireland/NIRE-23455D/2020\|EPI_ISL_ | .................... | .................... | .................... | .................... | .................... | .................... | |
| U | 6365 | hCoV-19/Northern Ireland/NIRE-23458A/2020\|EPI_ISL_ | .................... | .................... | .................... | .................... | .................... | .................... | |
| U | 6366 | hCoV-19/Northern Ireland/NIRE-2345A8/2020\|EPI_ISL_ | .................... | .................... | .................... | .................... | NNN................. | .................... | 1 |
| U | 6367 | hCoV-19/Northern Ireland/NIRE-2345F3/2020\|EPI_ISL_ | .................... | .................... | .................... | .................... | NNN................. | .................... | 1 |
| U | 6372 | hCoV-19/Northern Ireland/NIRE-2346D2/2020\|EPI_ISL_ | .................... | .................... | .................... | .................... | .................... | .................... | |
| U | 6373 | hCoV-19/Northern Ireland/NIRE-2346F0/2020\|EPI_ISL_ | .................... | .................... | .................... | .................... | .................... | .................... | |
| U | 6377 | hCoV-19/Chile/MA-CADIUMAG-17/2020\|EPI_ISL_681683\|2 | .................... | .................... | .................... | .................... | .................... | .................... | |
| U | 6378 | hCoV-19/Chile/MA-CADIUMAG-38/2020\|EPI_ISL_681684\|2 | .................... | .................... | .................... | .................... | .................... | .................... | |
| U | 6379 | hCoV-19/Chile/MA-CADIUMAG-26/2020\|EPI_ISL_681689\|2 | .................... | .................... | .................... | .................... | .................... | .................... | |
| U | 6380 | hCoV-19/Chile/MA-CADIUMAG-24/2020\|EPI_ISL_681690\|2 | .................... | .................... | .................... | .................... | .................... | .................... | |
| U | 6381 | hCoV-19/Chile/MA-CADIUMAG-27/2020\|EPI_ISL_681692\|2 | .................... | .................... | .................... | .................... | .................... | .................... | |
| U | 6382 | hCoV-19/Chile/MA-CADIUMAG-30/2020\|EPI_ISL_681695\|2 | .................... | .................... | .................... | .................... | .................... | .................... | |
| U | 6383 | hCoV-19/Chile/MA-CADIUMAG-31/2020\|EPI_ISL_681696\|2 | .................... | .................... | .................... | .................... | .................... | .................... | |
| U | 6384 | hCoV-19/Chile/MA-CADIUMAG-32/2020\|EPI_ISL_681697\|2 | .................... | .................... | .................... | .................... | .................... | .................... | |
| U | 6385 | hCoV-19/Chile/MA-CADIUMAG-33/2020\|EPI_ISL_681698\|2 | .................... | .................... | .................... | .................... | .................... | .................... | |
| U | 6386 | hCoV-19/Ireland/LK-UHL12/2020\|EPI_ISL_681721\|2020- | .................... | .................... | .................... | .................... | .................... | .................... | |
| U | 6387 | hCoV-19/Ireland/LK-UHL14/2020\|EPI_ISL_681731\|2020- | .................... | .................... | .................... | .................... | .................... | .................... | |
| U | 6388 | hCoV-19/Ireland/LK-UHL144/2020\|EPI_ISL_681734\|2020 | .................... | .................... | .................... | .................... | .................... | .................... | |
| U | 6389 | hCoV-19/Ireland/LK-UHL15/2020\|EPI_ISL_681736\|2020- | .................... | .................... | .................... | .................... | .................... | .................... | |
| U | 6390 | hCoV-19/Ireland/LK-UHL18/2020\|EPI_ISL_681737\|2020- | .................... | .................... | .................... | .................... | .................... | .................... | |
| U | 6391 | hCoV-19/Ireland/LK-UHL28/2020\|EPI_ISL_681739\|2020- | .................... | .................... | .................... | .................... | .................... | .................... | |
| U | 6392 | hCoV-19/Ireland/LK-UHL294/2020\|EPI_ISL_681742\|2020 | .................... | .................... | .................... | .................... | .................... | .................... | |
| U | 6393 | hCoV-19/Ireland/LK-UHL30/2020\|EPI_ISL_681744\|2020- | .................... | .................... | .................... | .................... | .................... | .................... | |
| U | 6394 | hCoV-19/Ireland/LK-UHL307/2020\|EPI_ISL_681747\|2020 | .................... | .................... | .................... | .................... | .................... | .................... | |
| U | 6395 | hCoV-19/Ireland/LK-UHL308/2020\|EPI_ISL_681748\|2020 | .................... | .................... | .................... | .................... | .................... | .................... | |
| U | 6396 | hCoV-19/Ireland/LK-UHL31/2020\|EPI_ISL_681749\|2020- | .................... | .................... | .................... | .................... | .................... | .................... | |
| U | 6397 | hCoV-19/Ireland/LK-UHL4/2020\|EPI_ISL_681752\|2020-0 | .................... | .................... | .................... | .................... | .................... | .................... | |
| U | 6398 | hCoV-19/Ireland/LK-UHL53/2020\|EPI_ISL_681756\|2020- | .................... | .................... | .................... | .................... | .................... | .................... | |
| U | 6399 | hCoV-19/Ireland/LK-UHL6/2020\|EPI_ISL_681757\|2020-0 | .................... | .................... | .................... | .................... | .................... | .................... | |
| U | 6400 | hCoV-19/Ireland/LK-UHL7/2020\|EPI_ISL_681759\|2020-0 | .................... | .................... | .................... | .................... | .................... | .................... | |
| U | 6401 | hCoV-19/England/CAMC-B09F96/2020\|EPI_ISL_673148\|20 | .................... | .................... | .................... | .................... | .................... | .................... | |
| U | 6402 | hCoV-19/England/CAMC-B0941F/2020\|EPI_ISL_673149\|20 | .................... | .................... | .................... | .................... | .................... | .................... | |
| U | 6403 | hCoV-19/England/CAMC-B09664/2020\|EPI_ISL_673150\|20 | .................... | .................... | .................... | .................... | .................... | .................... | |
| U | 6404 | hCoV-19/England/CAMC-B098D7/2020\|EPI_ISL_673151\|20 | .................... | .................... | .................... | .................... | .................... | .................... | |
| U | 6405 | hCoV-19/England/CAMC-B0A469/2020\|EPI_ISL_673152\|20 | .................... | .A.................. | .................... | .................... | ..........T......... | .................... | |
| U | 6406 | hCoV-19/England/CAMC-B0A557/2020\|EPI_ISL_673153\|20 | .................... | .................... | .................... | .................... | .................... | .................... | |
| U | 6407 | hCoV-19/England/CAMC-B09594/2020\|EPI_ISL_673154\|20 | .................... | .A.............G.... | .................... | .................... | .................... | ..A................. | |
| U | 6408 | hCoV-19/England/CAMC-B098F5/2020\|EPI_ISL_673155\|20 | .................... | .................... | .................... | .................... | .................... | .................... | |
| U | 6409 | hCoV-19/England/CAMC-B097CB/2020\|EPI_ISL_673156\|20 | .................... | .................... | .................... | .................... | .................... | .................... | |
| U | 6410 | hCoV-19/England/CAMC-B09831/2020\|EPI_ISL_673157\|20 | .................... | .................... | .................... | .................... | .................... | .................... | |
| U | 6411 | hCoV-19/England/CAMC-B0953A/2020\|EPI_ISL_673158\|20 | .................... | .................... | .................... | .................... | .................... | .................... | |
| U | 6412 | hCoV-19/England/CAMC-B0A487/2020\|EPI_ISL_673159\|20 | .................... | .................... | .................... | .................... | .................... | .................... | |
| U | 6413 | hCoV-19/England/CAMC-B0A01A/2020\|EPI_ISL_673160\|20 | .................... | .................... | .................... | .................... | .................... | .................... | |
| U | 6414 | hCoV-19/England/CAMC-B09DBA/2020\|EPI_ISL_673161\|20 | .................... | .A.................. | .................... | .................... | .................... | ..A................. | |
| U | 6415 | hCoV-19/England/CAMC-B09A4A/2020\|EPI_ISL_673162\|20 | .................... | .................... | .................... | .................... | .................... | .................... | |
| U | 6416 | hCoV-19/England/CAMC-B0A539/2020\|EPI_ISL_673163\|20 | .................... | .................... | .................... | .................... | .................... | .................... | |
| U | 6417 | hCoV-19/England/CAMC-B08F1F/2020\|EPI_ISL_673164\|20 | .................... | .................... | .................... | .................... | ......T............. | .................... | |
| U | 6418 | hCoV-19/England/CAMC-B09E7B/2020\|EPI_ISL_673165\|20 | .................... | .................... | .................... | .................... | .................... | .................... | |
| U | 6419 | hCoV-19/England/CAMC-B0A584/2020\|EPI_ISL_673166\|20 | .................... | .................... | .................... | .................... | .................... | .................... | |
| U | 6420 | hCoV-19/England/CAMC-B09C26/2020\|EPI_ISL_673167\|20 | .................... | .................... | .................... | .................... | .................... | .................... | |
| U | 6421 | hCoV-19/England/CAMC-B09B1A/2020\|EPI_ISL_673168\|20 | .................... | .................... | .................... | .................... | .................... | .................... | |
| U | 6422 | hCoV-19/England/CAMC-B08B66/2020\|EPI_ISL_673169\|20 | .................... | .................... | .................... | .................... | .................... | .................... | |
| U | 6423 | hCoV-19/England/CAMC-B09734/2020\|EPI_ISL_673170\|20 | .................... | .................... | .................... | .................... | .................... | .................... | |
| U | 6424 | hCoV-19/England/CAMC-B098E6/2020\|EPI_ISL_673171\|20 | .................... | .................... | .................... | .................... | .................... | .................... | |
| U | 6425 | hCoV-19/England/CAMC-B09172/2020\|EPI_ISL_673172\|20 | .................... | .................... | .................... | .................... | .................... | .................... | |
| U | 6426 | hCoV-19/England/CAMC-B0A171/2020\|EPI_ISL_673173\|20 | .................... | .................... | .................... | .................... | .................... | .................... | |
| U | 6427 | hCoV-19/England/CAMC-B09A77/2020\|EPI_ISL_673175\|20 | .................... | .................... | .................... | .................... | .................... | .................... | |
| U | 6428 | hCoV-19/England/CAMC-B08FA6/2020\|EPI_ISL_673176\|20 | .................... | .................... | .................... | .................... | .................... | .................... | |
| U | 6429 | hCoV-19/England/CAMC-B09A3B/2020\|EPI_ISL_673177\|20 | .................... | .................... | .................... | .................... | .................... | .................... | |
| U | 6430 | hCoV-19/England/CAMC-B08D8E/2020\|EPI_ISL_673178\|20 | .................... | .................... | .................... | .................... | .................... | .................... | |
| U | 6431 | hCoV-19/England/CAMC-B095FE/2020\|EPI_ISL_673179\|20 | .................... | .................... | .................... | .................... | .................... | .................... | |
| U | 6432 | hCoV-19/England/CAMC-B0936D/2020\|EPI_ISL_673180\|20 | .................... | .................... | .................... | .................... | .................... | .................... | |
| U | 6433 | hCoV-19/England/CAMC-B090DF/2020\|EPI_ISL_673182\|20 | .................... | .................... | .................... | .................... | .................... | .................... | |
| U | 6434 | hCoV-19/England/CAMC-B09D6F/2020\|EPI_ISL_673183\|20 | .................... | .................... | .................... | .................... | .................... | .................... | |
| U | 6435 | hCoV-19/England/CAMC-B09770/2020\|EPI_ISL_673184\|20 | .................... | .................... | .................... | .................... | .................... | .................... | |
| U | 6436 | hCoV-19/England/CAMC-B09CEA/2020\|EPI_ISL_673185\|20 | .................... | .................... | .................... | .................... | .................... | .................... | |
| U | 6437 | hCoV-19/England/CAMC-B0995C/2020\|EPI_ISL_673186\|20 | .................... | .................... | .................... | .................... | .................... | .................... | |
| U | 6438 | hCoV-19/England/CAMC-B098AA/2020\|EPI_ISL_673187\|20 | .................... | .................... | .................... | .................... | .................... | .................... | |

U 6439 hCoV-19/England/CAMC-B09B92/2020|EPI_ISL_673188|20 ···················· ···················· ····················· ···················· ···················· ····················
U 6440 hCoV-19/England/CAMC-B090EE/2020|EPI_ISL_673189|20 ···················· ···················· ····················· ···················· ···················· ····················
U 6441 hCoV-19/England/CAMC-B09488/2020|EPI_ISL_673190|20 ···················· ···················· ····················· ···················· ···················· ····················
U 6442 hCoV-19/England/CAMC-B09C80/2020|EPI_ISL_673191|20 ···················· ···················· ····················· ···················· ···················· ····················
U 6443 hCoV-19/England/CAMC-B09761/2020|EPI_ISL_673192|20 ···················· ···················· ····················· ···················· ···················· ····················
U 6444 hCoV-19/England/CAMC-B09725/2020|EPI_ISL_673193|20 ···················· ···················· ····················· ···················· ···················· ····················
U 6445 hCoV-19/England/CAMC-B0A52A/2020|EPI_ISL_673194|20 ···················· ···················· ····················· ···················· ······T············· ····················
U 6446 hCoV-19/England/CAMC-B0A5EE/2020|EPI_ISL_673195|20 ···················· ···················· ····················· ···················· ···················· ····················
U 6447 hCoV-19/England/CAMC-B097BC/2020|EPI_ISL_673196|20 ···················· ···················· ····················· ···················· ···················· ····················
U 6448 hCoV-19/England/CAMC-B08FD3/2020|EPI_ISL_673197|20 ···················· ···················· ····················· ···················· ···················· ····················
U 6449 hCoV-19/England/CAMC-B08E8B/2020|EPI_ISL_673198|20 ···················· ···················· ····················· ···················· ···················· ····················
U 6450 hCoV-19/England/CAMC-B09A2C/2020|EPI_ISL_673199|20 ···················· ···················· ····················· ···················· ···················· ····················
U 6451 hCoV-19/England/CAMC-B09804/2020|EPI_ISL_673200|20 ···················· ···················· ····················· ···················· ···················· ····················
U 6452 hCoV-19/England/CAMC-B097DA/2020|EPI_ISL_673201|20 ···················· ···················· ····················· ···················· ···················· ····················
U 6453 hCoV-19/England/CAMC-B098C8/2020|EPI_ISL_673202|20 ···················· ···················· ····················· ···················· ···················· ···A················
U 6454 hCoV-19/England/CAMC-B0996B/2020|EPI_ISL_673203|20 ···················· ···················· ····················· ···················· ···················· ····················
U 6455 hCoV-19/England/CAMC-B0A50C/2020|EPI_ISL_673204|20 ···················· ···················· ····················· ···················· ···················· ····················
U 6456 hCoV-19/England/CAMC-B08D33/2020|EPI_ISL_673205|20 ···················· ·A·················· ····················· ···················· ···················· ···A················
U 6457 hCoV-19/England/CAMC-B09AD1/2020|EPI_ISL_673206|20 ···················· ···················· ····················· ···················· ···················· ····················
U 6458 hCoV-19/England/CAMC-B09FE1/2020|EPI_ISL_673207|20 ···················· ···················· ····················· ···················· ···················· ····················
U 6459 hCoV-19/England/CAMC-B0A3F3/2020|EPI_ISL_673208|20 ···················· ···················· ····················· ···················· ···················· ····················
U 6460 hCoV-19/England/CAMC-B09CAE/2020|EPI_ISL_673209|20 ···················· ···················· ····················· ···················· ···················· ····················
U 6461 hCoV-19/England/CAMC-B09A59/2020|EPI_ISL_673210|20 ···················· ···················· ····················· ···················· ···················· ····················
U 6462 hCoV-19/England/CAMC-B09CCC/2020|EPI_ISL_673211|20 ···················· ···················· ····················· ···················· ···················· ····················
U 6463 hCoV-19/England/CAMC-B08DAC/2020|EPI_ISL_673212|20 ···················· ···················· ····················· ···················· ···················· ····················
U 6464 hCoV-19/England/CAMC-B09154/2020|EPI_ISL_673213|20 ···················· ···················· ····················· ···················· ···················· ····················
U 6465 hCoV-19/England/CAMC-B08F2E/2020|EPI_ISL_673214|20 ···················· ···················· ····················· ···················· ···················· ····················
U 6466 hCoV-19/England/CAMC-B09E8A/2020|EPI_ISL_673215|20 ···················· ···················· ····················· ···················· ······T············· ····················
U 6467 hCoV-19/England/CAMC-B08E21/2020|EPI_ISL_673216|20 ···················· ···················· ····················· ···················· ···················· ····················
U 6468 hCoV-19/England/CAMC-B0985F/2020|EPI_ISL_673217|20 ···················· ···················· ····················· ···················· ···················· ····················
U 6469 hCoV-19/England/CAMC-B09ED5/2020|EPI_ISL_673219|20 ···················· ···················· ····················· ···················· ···················· ····················
U 6470 hCoV-19/England/CAMC-B09048/2020|EPI_ISL_673220|20 ···················· ···················· ····················· ···················· ···················· ····················
U 6471 hCoV-19/England/CAMC-B09D14/2020|EPI_ISL_673221|20 ···················· ···················· ····················· ···················· ···················· ····················
U 6472 hCoV-19/England/CAMC-B0943D/2020|EPI_ISL_673222|20 ···················· ···················· ····················· ···················· ···················· ····················
U 6473 hCoV-19/England/CAMC-B08E5E/2020|EPI_ISL_673223|20 ···················· ·A·················· ····················· ···················· ···················· ···A················
U 6474 hCoV-19/England/CAMC-B09BED/2020|EPI_ISL_673230|20 ···················· ···················· ····················· ···················· ···················· ····················
U 6475 hCoV-19/England/CAMC-B09FF0/2020|EPI_ISL_673232|20 ···················· ···················· ····················· ···················· ····T··············· ····················
U 6476 hCoV-19/England/ALDP-B840AF/2020|EPI_ISL_674478|20 ···················· ···················· ····················· ···················· ······T············· ····················
U 6477 hCoV-19/England/MILK-B88481/2020|EPI_ISL_674480|20 ···················· ···················· ····················· ···················· ···················· ····················
U 6478 hCoV-19/England/MILK-B887A6/2020|EPI_ISL_674481|20 ···················· ···················· ····················· ···················· ···················· ····················
U 6479 hCoV-19/England/MILK-B8C20B/2020|EPI_ISL_674482|20 ···················· ···················· ····················· ···················· ···················· ····················
U 6480 hCoV-19/England/MILK-B88384/2020|EPI_ISL_674483|20 ···················· ···················· ····················· ···················· ···················· ····················
U 6481 hCoV-19/England/ALDP-B84D11/2020|EPI_ISL_674484|20 ···················· ···················· ····················· ···················· ···················· ····················
U 6482 hCoV-19/England/ALDP-B854FD/2020|EPI_ISL_674485|20 ···················· ···················· ····················· ···················· ···················· ····················
U 6483 hCoV-19/England/ALDP-B84A29/2020|EPI_ISL_674486|20 ···················· ···················· ····················· ···················· ···················· ····················
U 6484 hCoV-19/England/ALDP-B83978/2020|EPI_ISL_674487|20 ···················· ···················· ····················· ···················· ···················· ····················
U 6485 hCoV-19/England/ALDP-B84AA1/2020|EPI_ISL_674488|20 ···················· ···················· ····················· ···················· ···················· ····················
U 6486 hCoV-19/England/ALDP-B84C41/2020|EPI_ISL_674490|20 ···················· ···················· ····················· ···················· ···················· ····················
U 6487 hCoV-19/England/MILK-B8C1D1/2020|EPI_ISL_674491|20 ···················· ···················· ····················· ···················· ···················· ····················
U 6488 hCoV-19/England/MILK-B8C0F2/2020|EPI_ISL_674492|20 ···················· ···················· ····················· ···················· ···················· ····················
U 6489 hCoV-19/England/ALDP-B8443A/2020|EPI_ISL_674493|20 ···················· ···················· ····················· ···················· ···················· ····················
U 6490 hCoV-19/England/MILK-B8C0C5/2020|EPI_ISL_674494|20 ···················· ···················· ····················· ···················· ···················· ····················
U 6491 hCoV-19/England/ALDP-B83F2B/2020|EPI_ISL_674495|20 ···················· ···················· ····················· ···················· ···················· ····················
U 6492 hCoV-19/England/MILK-B8F411/2020|EPI_ISL_674496|20 ···················· ···················· ····················· ···················· ···················· ····················
U 6493 hCoV-19/England/MILK-B8F6A2/2020|EPI_ISL_674497|20 ···················· ···················· ····················· ···················· ···················· ····················
U 6494 hCoV-19/England/QEUH-B7D71D/2020|EPI_ISL_674498|20 ···················· ···················· ····················· ···················· ···················· ····················
U 6495 hCoV-19/England/QEUH-B7F8CD/2020|EPI_ISL_674499|20 ···················· ···················· ····················· ···················· ···················· ····················
U 6496 hCoV-19/England/MILK-B8ECDE/2020|EPI_ISL_674511|20 ···················· ···················· ····················· ···················· ···················· ····················
U 6497 hCoV-19/England/MILK-B8E9F5/2020|EPI_ISL_674512|20 ···················· ···················· ····················· ···················· ···················· ····················
U 6498 hCoV-19/England/MILK-B8E3F7/2020|EPI_ISL_674513|20 ···················· ···················· ····················· ···················· ···················· ····················
U 6499 hCoV-19/England/QEUH-B7CE28/2020|EPI_ISL_674514|20 ···················· ···················· ····················· ···················· ···················· ····················
U 6500 hCoV-19/England/MILK-B8DD45/2020|EPI_ISL_674515|20 ···················· ···················· ····················· ···················· ······T············· ····················
U 6501 hCoV-19/England/QEUH-B7F520/2020|EPI_ISL_674516|20 ···················· ···················· ····················· ···················· ···················· ····················
U 6502 hCoV-19/England/MILK-B8E8CB/2020|EPI_ISL_674517|20 ···················· ···················· ····················· ···················· ···················· ····················
U 6503 hCoV-19/England/MILK-B8E430/2020|EPI_ISL_674519|20 ···················· ···················· ····················· ···················· ···················· ····················
U 6504 hCoV-19/England/MILK-B8ED62/2020|EPI_ISL_674521|20 ···················· ···················· ····················· ···················· ···················· ····················
U 6505 hCoV-19/England/QEUH-B80999/2020|EPI_ISL_674522|20 ···················· ···················· ····················· ···················· ···················· ····················
U 6506 hCoV-19/England/MILK-B8E4B8/2020|EPI_ISL_674523|20 ···················· ·A·················· ····················· ···················· ···················· ···A················
U 6507 hCoV-19/England/MILK-B8E02D/2020|EPI_ISL_674524|20 ···················· ···················· ····················· ···················· ···················· ····················
U 6508 hCoV-19/England/MILK-B8EBB3/2020|EPI_ISL_674525|20 ···················· ···················· ····················· ···················· ···················· ····················
U 6509 hCoV-19/England/MILK-B8F7BE/2020|EPI_ISL_674526|20 ···················· ···················· ····················· ···················· ···················· ····················
U 6510 hCoV-19/England/QEUH-B7F53F/2020|EPI_ISL_674527|20 ···················· ···················· ····················· ···················· ···················· ····················
U 6511 hCoV-19/England/QEUH-B7D72C/2020|EPI_ISL_674528|20 ···················· ···················· ····················· ···················· ···················· ····················
U 6512 hCoV-19/England/MILK-B8EC38/2020|EPI_ISL_674529|20 ···················· ···················· ····················· ···················· ···················· ····················
U 6513 hCoV-19/England/QEUH-B7D66B/2020|EPI_ISL_674530|20 ···················· ···················· ····················· ···················· ···················· ····················
U 6514 hCoV-19/England/MILK-B8F842/2020|EPI_ISL_674531|20 ···················· ·A·················· ····················· ···················· ···················· ···A················
U 6515 hCoV-19/England/MILK-B8DC66/2020|EPI_ISL_674532|20 ···················· ···················· ····················· ···················· ···················· ····················
U 6516 hCoV-19/England/MILK-B8EBD1/2020|EPI_ISL_674533|20 ···················· ·A·················· ····················· ···················· ···················· ····················
U 6517 hCoV-19/England/MILK-B8F4C6/2020|EPI_ISL_674534|20 ···················· ···················· ····················· ···················· ······T············· ····················
U 6518 hCoV-19/England/MILK-B8F7DC/2020|EPI_ISL_674535|20 ···················· ·A·················· ····················· ···················· ···················· ····················

| | | | | | | | |
|---|---|---|---|---|---|---|---|
| U | 6519 hCoV-19/England/MILK-B8F51E/2020\|EPI_ISL_674536\|20 | ···················· | ···················· | ···················· | ···················· | ···················· | ···················· |
| U | 6520 hCoV-19/England/MILK-B8EB0E/2020\|EPI_ISL_674538\|20 | ···················· | ···················· | ···················· | ···················· | ···················· | ···················· |
| U | 6521 hCoV-19/England/MILK-B8E97D/2020\|EPI_ISL_674539\|20 | ···················· | ···················· | ···················· | ···················· | ···················· | ···················· |
| U | 6522 hCoV-19/England/MILK-B8EB68/2020\|EPI_ISL_674540\|20 | ···················· | ···················· | ···················· | ···················· | ···················· | ···················· |
| U | 6523 hCoV-19/England/MILK-B8F639/2020\|EPI_ISL_674541\|20 | ···················· | ···················· | ···················· | ···················· | ···················· | ···················· |
| U | 6524 hCoV-19/England/MILK-B8F9F4/2020\|EPI_ISL_674542\|20 | ···················· | ···················· | ···················· | ···················· | ···················· | ···················· |
| U | 6525 hCoV-19/England/QEUH-B7D64D/2020\|EPI_ISL_674543\|20 | ···················· | ···················· | ···················· | ···················· | ···················· | ···················· |
| U | 6526 hCoV-19/England/QEUH-B7CF25/2020\|EPI_ISL_674544\|20 | ···················· | ···················· | ···················· | ···················· | ···················· | ···················· |
| U | 6527 hCoV-19/England/QEUH-B7F4C9/2020\|EPI_ISL_674545\|20 | ···················· | ···················· | ···················· | ···················· | ···················· | ···················· |
| U | 6528 hCoV-19/England/MILK-B8F89D/2020\|EPI_ISL_674547\|20 | ···················· | ···················· | ···················· | ···················· | ···················· | ···················· |
| U | 6529 hCoV-19/England/MILK-B8F815/2020\|EPI_ISL_674548\|20 | ···················· | ···················· | ···················· | ···················· | ···················· | ···················· |
| U | 6530 hCoV-19/England/MILK-B8F46C/2020\|EPI_ISL_674549\|20 | ···················· | ···················· | ···················· | ···················· | ···················· | ···················· |
| U | 6531 hCoV-19/England/QEUH-B7CFAD/2020\|EPI_ISL_674550\|20 | ···················· | ···················· | ···················· | ···················· | ···················· | ···················· |
| U | 6532 hCoV-19/England/MILK-B8DD18/2020\|EPI_ISL_674553\|20 | ···················· | ···················· | ···················· | ···················· | ···················· | ···················· |
| U | 6533 hCoV-19/England/MILK-B8F8E8/2020\|EPI_ISL_674554\|20 | ···················· | ···················· | ···················· | ···················· | ···················· | ···················· |
| U | 6534 hCoV-19/England/MILK-B8F52D/2020\|EPI_ISL_674560\|20 | ···················· | ···················· | ···················· | ···················· | ···················· | ···················· |
| U | 6535 hCoV-19/England/MILK-B8EE9C/2020\|EPI_ISL_674561\|20 | ···················· | ·A·················· | ···················· | ···················· | ···················· | ···················· |
| U | 6536 hCoV-19/England/MILK-B8DCC0/2020\|EPI_ISL_674562\|20 | ···················· | ···················· | ···················· | ···················· | ···················· | ···················· |
| U | 6537 hCoV-19/England/MILK-B88928/2020\|EPI_ISL_675843\|20 | ···················· | ···················· | ···················· | ···················· | ···················· | ···················· |
| U | 6538 hCoV-19/England/MILK-B889A0/2020\|EPI_ISL_675844\|20 | ···················· | ···················· | ···················· | ···················· | ···················· | ···················· |
| U | 6539 hCoV-19/England/ALDP-B85994/2020\|EPI_ISL_675847\|20 | ···················· | ···················· | ···················· | ···················· | ···················· | ···················· |
| U | 6540 hCoV-19/England/MILK-B8C98E/2020\|EPI_ISL_675849\|20 | ···················· | ···················· | ···················· | ···················· | ···················· | ···················· |
| U | 6541 hCoV-19/England/MILK-B8C7EE/2020\|EPI_ISL_675850\|20 | ···················· | ···················· | ···················· | ···················· | ···················· | ···················· |
| U | 6542 hCoV-19/England/MILK-B88B04/2020\|EPI_ISL_675851\|20 | ···················· | ···················· | ···················· | ···················· | ···················· | ···················· |
| U | 6543 hCoV-19/England/ALDP-B84E96/2020\|EPI_ISL_675852\|20 | ···················· | ···················· | ···················· | ···················· | ···················· | ···················· |
| U | 6544 hCoV-19/England/MILK-B897F0/2020\|EPI_ISL_675853\|20 | ···················· | ···················· | ···················· | ···················· | ···················· | ···················· |
| U | 6545 hCoV-19/England/MILK-B88CB6/2020\|EPI_ISL_675854\|20 | ···················· | ···················· | ···················· | ···················· | ···················· | ···················· |
| U | 6546 hCoV-19/England/ALDP-B859B2/2020\|EPI_ISL_675855\|20 | ···················· | ···················· | ···················· | ···················· | ···················· | ···················· |
| U | 6547 hCoV-19/England/MILK-B88ABC/2020\|EPI_ISL_675857\|20 | ···················· | ···················· | ···················· | ···················· | ···················· | ···················· |
| U | 6548 hCoV-19/England/MILK-B896E4/2020\|EPI_ISL_675859\|20 | ···················· | ···················· | ···················· | ···················· | ···················· | ···················· |
| U | 6549 hCoV-19/England/MILK-B88E92/2020\|EPI_ISL_675862\|20 | ···················· | ···················· | ···················· | ···················· | ···················· | ···················· |
| U | 6550 hCoV-19/England/MILK-B8875B/2020\|EPI_ISL_675863\|20 | ···················· | ···················· | ···················· | ···················· | ···················· | ···················· |
| U | 6551 hCoV-19/England/MILK-B8972D/2020\|EPI_ISL_675865\|20 | ···················· | ···················· | ···················· | ···················· | ···················· | ···················· |
| U | 6552 hCoV-19/England/MILK-B8CB5B/2020\|EPI_ISL_675866\|20 | ···················· | ···················· | ···················· | ···················· | ···················· | ···················· |
| U | 6553 hCoV-19/England/MILK-B88A61/2020\|EPI_ISL_675867\|20 | ···················· | ···················· | ···················· | ···················· | ···················· | ···················· |
| U | 6554 hCoV-19/England/ALDP-B84EB4/2020\|EPI_ISL_675868\|20 | ···················· | ···················· | ···················· | ···················· | ···················· | ···················· |
| U | 6555 hCoV-19/England/MILK-B8CAF4/2020\|EPI_ISL_675871\|20 | ···················· | ···················· | ···················· | ···················· | ···················· | ···················· |
| U | 6556 hCoV-19/England/MILK-B8C748/2020\|EPI_ISL_675872\|20 | ···················· | ···················· | ···················· | ···················· | ···················· | ···················· |
| U | 6557 hCoV-19/England/MILK-B89550/2020\|EPI_ISL_675873\|20 | ···················· | ···················· | ···················· | ···················· | ···················· | ···················· |
| U | 6558 hCoV-19/England/MILK-B8C9D9/2020\|EPI_ISL_675874\|20 | ···················· | ···················· | ···················· | ···················· | ···················· | ···················· |
| U | 6559 hCoV-19/England/MILK-B8C7B1/2020\|EPI_ISL_675876\|20 | ···················· | ···················· | ···················· | ···················· | ···················· | ···················· |
| U | 6560 hCoV-19/England/MILK-B88CE3/2020\|EPI_ISL_675877\|20 | ···················· | ···················· | ···················· | ···················· | ···················· | ···················· |
| U | 6561 hCoV-19/England/ALDP-B84DC6/2020\|EPI_ISL_675878\|20 | ···················· | ···················· | ···················· | ···················· | ···················· | ···················· |
| U | 6562 hCoV-19/England/MILK-B8965D/2020\|EPI_ISL_675879\|20 | ···················· | ···················· | ···················· | ···················· | ···················· | ···················· |
| U | 6563 hCoV-19/England/MILK-B8980C/2020\|EPI_ISL_675881\|20 | ···················· | ···················· | ···················· | ···················· | ···················· | ···················· |
| U | 6564 hCoV-19/England/ALDP-B84DA8/2020\|EPI_ISL_675882\|20 | ···················· | ···················· | ···················· | ···················· | ···················· | ···················· |
| U | 6565 hCoV-19/England/MILK-B88D77/2020\|EPI_ISL_675884\|20 | ···················· | ···················· | ···················· | ···················· | ···················· | ···················· |
| U | 6566 hCoV-19/England/MILK-B88A52/2020\|EPI_ISL_675889\|20 | ···················· | ···················· | ···················· | ···················· | ···················· | ···················· |
| U | 6567 hCoV-19/Saudi Arabia/KAUST-MADINAH1362/2020\|EPI_IS | ···················· | ···················· | ···················· | ···················· | ···················· | ···················· |
| U | 6568 hCoV-19/Saudi Arabia/KAUST-MADINAH1363/2020\|EPI_IS | ···················· | ···················· | ···················· | ···················· | ···················· | ···················· |
| U | 6569 hCoV-19/Saudi Arabia/KAUST-MADINAH1366/2020\|EPI_IS | ···················· | ·A·················· | ···················· | ···················· | ···················· | ··A················· |
| U | 6570 hCoV-19/Saudi Arabia/KAUST-MADINAH1367/2020\|EPI_IS | ···················· | ·A·················· | ···················· | ···················· | ···················· | ··A················· |
| U | 6571 hCoV-19/Saudi Arabia/KAUST-MADINAH1372/2020\|EPI_IS | ···················· | ·A·················· | ···················· | ···················· | ···················· | ··A················· |
| U | 6572 hCoV-19/Saudi Arabia/KAUST-MADINAH1373/2020\|EPI_IS | ···················· | ···················· | ···················· | ···················· | ···················· | ···················· |
| U | 6573 hCoV-19/Saudi Arabia/KAUST-MADINAH1374/2020\|EPI_IS | ···················· | ···················· | ···················· | ···················· | ···················· | ···················· |
| U | 6574 hCoV-19/Saudi Arabia/KAUST-MADINAH1375/2020\|EPI_IS | ···················· | ···················· | ···················· | ···················· | ···················· | ···················· |
| U | 6575 hCoV-19/Saudi Arabia/KAUST-MADINAH1379/2020\|EPI_IS | ···················· | ···················· | ···················· | ···················· | ···················· | ···················· |
| U | 6576 hCoV-19/Saudi Arabia/KAUST-MADINAH1381/2020\|EPI_IS | ···················· | ·A·················· | ···················· | ···················· | ···················· | ··A················· |
| U | 6577 hCoV-19/Saudi Arabia/KAUST-MADINAH1478/2020\|EPI_IS | ···················· | ···················· | ···················· | ···················· | ···················· | ···················· |
| U | 6578 hCoV-19/Saudi Arabia/KAUST-MADINAH1480/2020\|EPI_IS | ···················· | ·A·················· | ···················· | ···················· | ···················· | ··A················· |
| U | 6579 hCoV-19/Saudi Arabia/KAUST-MADINAH1483/2020\|EPI_IS | ···················· | ···················· | ···················· | ···················· | ···················· | ···················· |
| U | 6580 hCoV-19/Saudi Arabia/KAUST-MADINAH1484/2020\|EPI_IS | ···················· | ·A·················· | ···················· | ···················· | ···················· | ··A················· |
| U | 6581 hCoV-19/Saudi Arabia/KAUST-MADINAH1487/2020\|EPI_IS | ···················· | ···················· | ···················· | ···················· | ···················· | ···················· |
| U | 6582 hCoV-19/Saudi Arabia/KAUST-MADINAH1488/2020\|EPI_IS | ···················· | ·A·················· | ···················· | ···················· | ···················· | ···················· |
| U | 6584 hCoV-19/Saudi Arabia/KAUST-MADINAH250/2020\|EPI_ISL | ···················· | ···················· | ···················· | ···················· | ···················· | ···················· |
| U | 6585 hCoV-19/Saudi Arabia/KAUST-MADINAH268/2020\|EPI_ISL | ···················· | ···················· | ···················· | ···················· | ···················· | ···················· |
| U | 6586 hCoV-19/Saudi Arabia/KAUST-MADINAH284/2020\|EPI_ISL | ···················· | ·A·················· | ···················· | ···················· | ···················· | ··A················· |
| U | 6587 hCoV-19/Saudi Arabia/KAUST-MADINAH451/2020\|EPI_ISL | ···················· | ···················· | ···················· | ···················· | ···················· | ···················· |
| U | 6588 hCoV-19/Saudi Arabia/KAUST-MADINAH453/2020\|EPI_ISL | ···················· | ···················· | ···················· | ···················· | ···················· | ···················· |
| U | 6589 hCoV-19/Saudi Arabia/KAUST-MADINAH602/2020\|EPI_ISL | ···················· | ···················· | ···················· | ···················· | ···················· | ···················· |
| U | 6590 hCoV-19/Saudi Arabia/KAUST-MADINAH636/2020\|EPI_ISL | ···················· | ···················· | ···················· | ···················· | ···················· | ···················· |
| U | 6591 hCoV-19/Saudi Arabia/KAUST-MADINAH654/2020\|EPI_ISL | ···················· | ·A·················· | ···················· | ···················· | ···················· | ··A················· |
| U | 6592 hCoV-19/Saudi Arabia/KAUST-MADINAH662/2020\|EPI_ISL | ···················· | ···················· | ···················· | ···················· | ···················· | ···················· |
| U | 6593 hCoV-19/Saudi Arabia/KAUST-MADINAH663/2020\|EPI_ISL | ···················· | ···················· | ···················· | ···················· | ···················· | ···················· |
| U | 6594 hCoV-19/Saudi Arabia/KAUST-MADINAH665/2020\|EPI_ISL | ···················· | ···················· | ···················· | ···················· | ···················· | ···················· |
| U | 6595 hCoV-19/Saudi Arabia/KAUST-MADINAH673/2020\|EPI_ISL | ···················· | ···················· | ···················· | ···················· | ···················· | ···················· |
| U | 6596 hCoV-19/England/EXET-139D42/2020\|EPI_ISL_679340\|20 | ···················· | ···················· | ···················· | ···················· | ···················· | ···················· |
| U | 6597 hCoV-19/England/EXET-139D60/2020\|EPI_ISL_679341\|20 | ···················· | ···················· | ···················· | ···················· | ······T············· | ···················· |
| U | 6598 hCoV-19/England/EXET-139DD9/2020\|EPI_ISL_679343\|20 | ···················· | ···················· | ···················· | ···················· | ···················· | ···················· |
| U | 6599 hCoV-19/England/EXET-139DE8/2020\|EPI_ISL_679344\|20 | ···················· | ···················· | ···················· | ···················· | ······T············· | ···················· |

| | | | | | | | | |
|---|---|---|---|---|---|---|---|---|
| U | 6600 | hCoV-19/England/EXET-139E03/2020\|EPI_ISL_679346\|20 | ···················· | ···················· | ···················· | ···················· | ······T············· | ···················· |
| U | 6601 | hCoV-19/England/EXET-139E8B/2020\|EPI_ISL_679348\|20 | ···················· | ···················· | ···················· | ···················· | ······T············· | ···················· |
| U | 6602 | hCoV-19/England/EXET-139E9A/2020\|EPI_ISL_679349\|20 | ···················· | ···················· | ···················· | ···················· | ···················· | ···················· |
| U | 6603 | hCoV-19/Northern Ireland/NIRE-233FC9/2020\|EPI_ISL_ | ···················· | ···················· | ···················· | ···················· | ···················· | ···················· |
| U | 6604 | hCoV-19/Northern Ireland/NIRE-233FF6/2020\|EPI_ISL_ | ···················· | ···················· | ···················· | ···················· | ···················· | ···················· |
| U | 6607 | hCoV-19/Northern Ireland/NIRE-23403E/2020\|EPI_ISL_ | ···················· | ···················· | ···················· | ···················· | ···················· | ···················· |
| U | 6608 | hCoV-19/Northern Ireland/NIRE-23404D/2020\|EPI_ISL_ | ···················· | ···················· | ···················· | ···················· | ···················· | ···················· |
| U | 6609 | hCoV-19/Northern Ireland/NIRE-23405C/2020\|EPI_ISL_ | ···················· | ···················· | ···················· | ···················· | ···················· | ···················· |
| U | 6611 | hCoV-19/Northern Ireland/NIRE-234089/2020\|EPI_ISL_ | ···················· | ···················· | ···················· | ···················· | ···················· | ···················· |
| U | 6612 | hCoV-19/Northern Ireland/NIRE-234098/2020\|EPI_ISL_ | ···················· | ···················· | ···················· | ···················· | ···················· | ···················· |
| U | 6613 | hCoV-19/Northern Ireland/NIRE-2340D4/2020\|EPI_ISL_ | ···················· | ···················· | ···················· | ···················· | ···················· | ···················· |
| U | 6615 | hCoV-19/Northern Ireland/NIRE-234168/2020\|EPI_ISL_ | ···················· | ···················· | ···················· | ···················· | ···················· | ···················· |
| U | 6616 | hCoV-19/Northern Ireland/NIRE-2341A4/2020\|EPI_ISL_ | ···················· | ···················· | ···················· | ···················· | ···················· | ···················· |
| U | 6617 | hCoV-19/Northern Ireland/NIRE-2341C2/2020\|EPI_ISL_ | ···················· | ···················· | ···················· | ···················· | ···················· | ···················· |
| U | 6618 | hCoV-19/England/MILK-B88E65/2020\|EPI_ISL_681660\|20 | ···················· | ···················· | ···················· | ···················· | ···················· | ···················· |
| U | 6619 | hCoV-19/England/ALDP-B85976/2020\|EPI_ISL_681661\|20 | ···················· | ···················· | ···················· | ···················· | ···················· | ···················· |
| U | 6620 | hCoV-19/England/ALDP-B853E1/2020\|EPI_ISL_681665\|20 | ···················· | ···················· | ···················· | ···················· | ···················· | ···················· |
| U | 6621 | hCoV-19/England/MILK-B8F921/2020\|EPI_ISL_681666\|20 | ···················· | ···················· | ···················· | ···················· | ···················· | ···················· |
| U | 6622 | hCoV-19/England/MILK-B961B6/2020\|EPI_ISL_681667\|20 | ···················· | ···················· | ···················· | ···················· | ···················· | ···················· |
| U | 6623 | hCoV-19/England/MILK-B96D2B/2020\|EPI_ISL_681668\|20 | ···················· | ···················· | ···················· | ···················· | ···················· | ···················· |
| U | 6624 | hCoV-19/England/MILK-B96FAD/2020\|EPI_ISL_681670\|20 | ···················· | ···················· | ···················· | ···················· | ···················· | ···················· |
| U | 6625 | hCoV-19/England/CAMC-B79B06/2020\|EPI_ISL_681671\|20 | ···················· | ···················· | ···················· | ···················· | ···················· | ···················· |
| U | 6626 | hCoV-19/England/CAMC-B79A27/2020\|EPI_ISL_681672\|20 | ···················· | ···················· | ···················· | ···················· | ···················· | ···················· |
| U | 6627 | hCoV-19/Chile/MA-CADIUMAG-1/2020\|EPI_ISL_681673\|20 | ···················· | ···················· | ···················· | ···················· | ···················· | ···················· |
| U | 6630 | hCoV-19/Chile/MA-CADIUMAG-15/2020\|EPI_ISL_681681\|2 | ···················· | ···················· | ···················· | ···················· | ···················· | ···················· |
| U | 6631 | hCoV-19/Chile/MA-CADIUMAG-16/2020\|EPI_ISL_681682\|2 | ···················· | ···················· | ···················· | ···················· | ···················· | ···················· |
| U | 6632 | hCoV-19/Denmark/DCGC-12719/2020\|EPI_ISL_682900\|202 | ···················· | ···················· | ···················· | ···················· | ···················· | ···················· |
| U | 6633 | hCoV-19/Denmark/DCGC-12720/2020\|EPI_ISL_682901\|202 | ···················· | ·A·················· | ···················· | ···················· | ···················· | ···················· |
| U | 6634 | hCoV-19/Denmark/DCGC-12721/2020\|EPI_ISL_682902\|202 | ···················· | ···················· | ···················· | ···················· | ···················· | ···················· |
| U | 6635 | hCoV-19/Denmark/DCGC-12722/2020\|EPI_ISL_682903\|202 | ···················· | ···················· | ···················· | ···················· | ···················· | ···················· |
| U | 6636 | hCoV-19/Denmark/DCGC-12723/2020\|EPI_ISL_682904\|202 | ···················· | ···················· | ···················· | ···················· | ···················· | ···················· |
| U | 6637 | hCoV-19/Denmark/DCGC-12724/2020\|EPI_ISL_682905\|202 | ···················· | ···················· | ···················· | ···················· | ···················· | ···················· |
| U | 6638 | hCoV-19/Denmark/DCGC-12726/2020\|EPI_ISL_682907\|202 | ···················· | ···················· | ···················· | ···················· | ···················· | ···················· |
| U | 6639 | hCoV-19/Denmark/DCGC-12727/2020\|EPI_ISL_682908\|202 | ···················· | ···················· | ···················· | ···················· | ···················· | ···················· |
| U | 6640 | hCoV-19/Denmark/DCGC-12729/2020\|EPI_ISL_682910\|202 | ···················· | ···················· | ···················· | ···················· | ······T············· | ···················· |
| U | 6641 | hCoV-19/Denmark/DCGC-12732/2020\|EPI_ISL_682913\|202 | ···················· | ···················· | ···················· | ···················· | ···················· | ···················· |
| U | 6642 | hCoV-19/Denmark/DCGC-12733/2020\|EPI_ISL_682914\|202 | ···················· | ···················· | ···················· | ···················· | ···················· | ···················· |
| U | 6643 | hCoV-19/Denmark/DCGC-12735/2020\|EPI_ISL_682916\|202 | ···················· | ···················· | ···················· | ···················· | ···················· | ···················· |
| U | 6644 | hCoV-19/Denmark/DCGC-12738/2020\|EPI_ISL_682919\|202 | ···················· | ···················· | ···················· | ···················· | ···················· | ···················· |
| U | 6645 | hCoV-19/Denmark/DCGC-12739/2020\|EPI_ISL_682920\|202 | ···················· | ···················· | ···················· | ···················· | ···················· | ···················· |
| U | 6646 | hCoV-19/Denmark/DCGC-12740/2020\|EPI_ISL_682921\|202 | ···················· | ···················· | ···················· | ···················· | ···················· | ···················· |
| U | 6647 | hCoV-19/Denmark/DCGC-12741/2020\|EPI_ISL_682922\|202 | ···················· | ···················· | ···················· | ···················· | ···················· | ···················· |
| U | 6648 | hCoV-19/Denmark/DCGC-12744/2020\|EPI_ISL_682925\|202 | ···················· | ···················· | ···················· | ···················· | ···················· | ···················· |
| U | 6649 | hCoV-19/Denmark/DCGC-12746/2020\|EPI_ISL_682927\|202 | ···················· | ···················· | ···················· | ···················· | ···················· | ···················· |
| U | 6650 | hCoV-19/Denmark/DCGC-12747/2020\|EPI_ISL_682928\|202 | ···················· | ···················· | ···················· | ···················· | ···················· | ···················· |
| U | 6651 | hCoV-19/Denmark/DCGC-12809/2020\|EPI_ISL_682990\|202 | ···················· | ·A·················· | ···················· | ···················· | ···················· | ···················· |
| U | 6652 | hCoV-19/Denmark/DCGC-12812/2020\|EPI_ISL_682993\|202 | ···················· | ···················· | ···················· | ···················· | ···················· | ···················· |
| U | 6653 | hCoV-19/Denmark/DCGC-12813/2020\|EPI_ISL_682994\|202 | ···················· | ···················· | ···················· | ···················· | ···················· | ···················· |
| U | 6654 | hCoV-19/Denmark/DCGC-12814/2020\|EPI_ISL_682995\|202 | ···················· | ···················· | ···················· | ···················· | ···················· | ···················· |
| U | 6655 | hCoV-19/Denmark/DCGC-12816/2020\|EPI_ISL_682997\|202 | ···················· | ···················· | ···················· | ···················· | ···················· | ···················· |
| U | 6656 | hCoV-19/Denmark/DCGC-12817/2020\|EPI_ISL_682998\|202 | ···················· | ···················· | ···················· | ···················· | ···················· | ···················· |
| U | 6657 | hCoV-19/England/CAMC-B0952B/2020\|EPI_ISL_673128\|20 | ···················· | ···················· | ···················· | ···················· | ···················· | ···················· |
| U | 6658 | hCoV-19/England/CAMC-B0A593/2020\|EPI_ISL_673129\|20 | ···················· | ···················· | ···················· | ···················· | ···················· | ···················· |
| U | 6659 | hCoV-19/England/CAMC-B0A047/2020\|EPI_ISL_673136\|20 | ···················· | ···················· | ···················· | ···················· | ···················· | ···················· |
| U | 6660 | hCoV-19/England/CAMC-B09822/2020\|EPI_ISL_673137\|20 | ···················· | ···················· | ···················· | ···················· | ···················· | ···················· |
| U | 6661 | hCoV-19/England/CAMC-B09A0E/2020\|EPI_ISL_673138\|20 | ···················· | ···················· | ···················· | ···················· | ···················· | ···················· |
| U | 6662 | hCoV-19/England/CAMC-B09FC3/2020\|EPI_ISL_673139\|20 | ···················· | ···················· | ···················· | ···················· | ···················· | ···················· |
| U | 6663 | hCoV-19/England/CAMC-B0A51B/2020\|EPI_ISL_673140\|20 | ···················· | ·A·················· | ···················· | ···················· | ···················· | ···A················ |
| U | 6664 | hCoV-19/England/CAMC-B0945B/2020\|EPI_ISL_673141\|20 | ···················· | ···················· | ···················· | ···················· | ···················· | ···················· |
| U | 6665 | hCoV-19/England/CAMC-B09F2D/2020\|EPI_ISL_673142\|20 | ···················· | ···················· | ···················· | ···················· | ···················· | ···················· |
| U | 6666 | hCoV-19/England/CAMC-B09F0F/2020\|EPI_ISL_673143\|20 | ···················· | ···················· | ···················· | ···················· | ···················· | ···················· |
| U | 6667 | hCoV-19/England/CAMC-B09CF9/2020\|EPI_ISL_673144\|20 | ···················· | ···················· | ···················· | ···················· | ···················· | ···················· |
| U | 6668 | hCoV-19/England/CAMC-B09D23/2020\|EPI_ISL_673146\|20 | ···················· | ···················· | ···················· | ···················· | ···················· | ···················· |
| U | 6669 | hCoV-19/England/ALDP-B84A38/2020\|EPI_ISL_674459\|20 | ···················· | ···················· | ···················· | ···················· | ···················· | ···················· |
| U | 6670 | hCoV-19/England/ALDP-B84F93/2020\|EPI_ISL_674467\|20 | ···················· | ···················· | ···················· | ···················· | ···················· | ···················· |
| U | 6671 | hCoV-19/England/MILK-B8C05C/2020\|EPI_ISL_674468\|20 | ···················· | ···················· | ···················· | ···················· | ···················· | ···················· |
| U | 6672 | hCoV-19/England/ALDP-B85299/2020\|EPI_ISL_674469\|20 | ···················· | ···················· | ···················· | ···················· | ···················· | ···················· |
| U | 6673 | hCoV-19/England/ALDP-B84449/2020\|EPI_ISL_674470\|20 | ···················· | ···················· | ···················· | ···················· | ···················· | ···················· |
| U | 6674 | hCoV-19/England/ALDP-B85439/2020\|EPI_ISL_674471\|20 | ···················· | ···················· | ···················· | ···················· | ···················· | ···················· |
| U | 6675 | hCoV-19/England/MILK-B882C3/2020\|EPI_ISL_674473\|20 | ···················· | ···················· | ···················· | ···················· | ···················· | ···················· |
| U | 6676 | hCoV-19/England/ALDP-B8442B/2020\|EPI_ISL_674474\|20 | ···················· | ·A·················· | ···················· | ···················· | ···················· | ···A················ |
| U | 6677 | hCoV-19/England/MILK-B88849/2020\|EPI_ISL_674475\|20 | ···················· | ···················· | ···················· | ···················· | ···················· | ···················· |
| U | 6678 | hCoV-19/England/ALDP-B83D21/2020\|EPI_ISL_674476\|20 | ···················· | ···················· | ···················· | ···················· | ······T············· | ···················· |
| U | 6679 | hCoV-19/England/ALDP-B83D12/2020\|EPI_ISL_674477\|20 | ···················· | ···················· | ···················· | ···················· | ···················· | ···················· |
| U | 6680 | hCoV-19/England/ALDP-B83BCD/2020\|EPI_ISL_675701\|20 | ···················· | ···················· | ···················· | ···················· | ···················· | ···················· |
| U | 6681 | hCoV-19/England/ALDP-B83204/2020\|EPI_ISL_675702\|20 | ···················· | ···················· | ···················· | ···················· | ···················· | ···················· |
| U | 6682 | hCoV-19/England/ALDP-B830BF/2020\|EPI_ISL_675703\|20 | ···················· | ···················· | ···················· | ···················· | ···················· | ···················· |
| U | 6683 | hCoV-19/England/ALDP-B8351A/2020\|EPI_ISL_675705\|20 | ···················· | ···················· | ···················· | ···················· | ···················· | ···················· |
| U | 6684 | hCoV-19/England/ALDP-B85B25/2020\|EPI_ISL_675706\|20 | ···················· | ·A·················· | ···················· | ···················· | ···················· | ···················· |
| U | 6685 | hCoV-19/England/ALDP-B82FE0/2020\|EPI_ISL_675707\|20 | ···················· | ···················· | ···················· | ···················· | ···················· | ···················· |

| | | | | | | | | |
|---|---|---|---|---|---|---|---|---|
| U | 6686 hCoV-19/England/ALDP-B83B63/2020\|EPI_ISL_675708\|20 | ·················· | ·················· | ·················· | ·················· | ·················· | ·················· | |
| U | 6687 hCoV-19/England/ALDP-B83116/2020\|EPI_ISL_675710\|20 | ·················· | ·A················ | ·················· | ·················· | ·················· | ··A··············· | |
| U | 6688 hCoV-19/England/MILK-B8C863/2020\|EPI_ISL_675711\|20 | ·················· | ·················· | ·················· | ·················· | ·················· | ·················· | |
| U | 6689 hCoV-19/England/MILK-B8981B/2020\|EPI_ISL_675712\|20 | ·················· | ·················· | ·················· | ·················· | ·················· | ·················· | |
| U | 6690 hCoV-19/England/MILK-B88A07/2020\|EPI_ISL_675713\|20 | ·················· | ·················· | ·················· | ·················· | ·················· | ·················· | |
| U | 6691 hCoV-19/England/ALDP-B84BAE/2020\|EPI_ISL_675715\|20 | ·················· | ·················· | ·················· | ·················· | ·····T············ | ·················· | |
| U | 6692 hCoV-19/England/MILK-B8C53F/2020\|EPI_ISL_675717\|20 | ·················· | ·················· | ·················· | ·················· | ·················· | ·················· | |
| U | 6693 hCoV-19/England/ALDP-B85A28/2020\|EPI_ISL_675720\|20 | ·················· | ·················· | ·················· | ·················· | ·················· | ·················· | |
| U | 6694 hCoV-19/England/MILK-B88CA7/2020\|EPI_ISL_675790\|20 | ·················· | ·················· | ·················· | ·················· | ·················· | ·················· | |
| U | 6695 hCoV-19/England/MILK-B8C696/2020\|EPI_ISL_675791\|20 | ·················· | ·················· | ·················· | ·················· | ·················· | ·················· | |
| U | 6696 hCoV-19/England/MILK-B8C8BE/2020\|EPI_ISL_675792\|20 | ·················· | ·················· | ·················· | ·················· | ·················· | ·················· | |
| U | 6697 hCoV-19/England/ALDP-B84704/2020\|EPI_ISL_675794\|20 | ·················· | ·················· | ·················· | ·················· | ·················· | ·················· | |
| U | 6698 hCoV-19/England/ALDP-B848A7/2020\|EPI_ISL_675798\|20 | ·················· | ·················· | ·················· | ·················· | ·················· | ·················· | |
| U | 6699 hCoV-19/Saudi Arabia/KAUST-MADINAH1148/2020\|EPI_IS | ·················· | ·A················ | ·················· | ·················· | ·················· | ··A··············· | |
| U | 6700 hCoV-19/Saudi Arabia/KAUST-MADINAH1149/2020\|EPI_IS | ·················· | ·A················ | ·················· | ·················· | ·················· | ··A··············· | |
| U | 6701 hCoV-19/Saudi Arabia/KAUST-MADINAH1150/2020\|EPI_IS | ·················· | ·A················ | ·················· | ·················· | ·················· | ··A··············· | |
| U | 6702 hCoV-19/Saudi Arabia/KAUST-MADINAH1151/2020\|EPI_IS | ·················· | ·················· | ·················· | ·················· | ·················· | ··A··············· | |
| U | 6703 hCoV-19/Saudi Arabia/KAUST-MADINAH1269/2020\|EPI_IS | ·················· | ·A················ | ·················· | ·················· | ·················· | ··A··············· | |
| U | 6704 hCoV-19/Saudi Arabia/KAUST-MADINAH1271/2020\|EPI_IS | ·················· | ·················· | ·················· | ·················· | ·················· | ·················· | |
| U | 6705 hCoV-19/Saudi Arabia/KAUST-MADINAH1274/2020\|EPI_IS | ·················· | ·A················ | ·················· | ·················· | ·················· | ··A··············· | |
| U | 6706 hCoV-19/Saudi Arabia/KAUST-MADINAH1277/2020\|EPI_IS | ·················· | ·················· | ·················· | ·················· | ·················· | ·················· | |
| U | 6707 hCoV-19/Saudi Arabia/KAUST-MADINAH1283/2020\|EPI_IS | ·················· | ·A················ | ·················· | ·················· | ·················· | ··A··············· | |
| U | 6708 hCoV-19/Saudi Arabia/KAUST-MADINAH1284/2020\|EPI_IS | ·················· | ·A················ | ·················· | ·················· | ·················· | ··A··············· | |
| U | 6709 hCoV-19/England/CAMC-BBC897/2020\|EPI_ISL_679251\|20 | ·················· | ·················· | ·················· | ·················· | ·················· | ·················· | |
| U | 6710 hCoV-19/England/CAMC-BBC90D/2020\|EPI_ISL_679255\|20 | ·················· | ·················· | ·················· | ·················· | ·················· | ·················· | |
| U | 6711 hCoV-19/England/CAMC-BBC976/2020\|EPI_ISL_679258\|20 | ·················· | ·················· | ·················· | ·················· | ·····T············ | ·················· | |
| U | 6712 hCoV-19/England/CAMC-BBC9B2/2020\|EPI_ISL_679261\|20 | ·················· | ·················· | ·················· | ·················· | ·················· | ·················· | |
| U | 6713 hCoV-19/England/CAMC-BBCA0A/2020\|EPI_ISL_679265\|20 | ·················· | ·················· | ·················· | ·················· | ·················· | ·················· | |
| U | 6714 hCoV-19/England/CAMC-BBCA19/2020\|EPI_ISL_679266\|20 | ·················· | ·················· | ·················· | ·················· | ·················· | ·················· | |
| U | 6715 hCoV-19/England/CAMC-BBCC31/2020\|EPI_ISL_679269\|20 | ·················· | ·················· | ·················· | ·················· | ·················· | ·················· | |
| U | 6716 hCoV-19/England/CAMC-BBCC40/2020\|EPI_ISL_679270\|20 | ·················· | ·················· | ·················· | ·················· | ·················· | ·················· | |
| U | 6717 hCoV-19/England/CAMC-BBCCF5/2020\|EPI_ISL_679275\|20 | ·················· | ·················· | ·················· | ·················· | ·················· | ·················· | |
| U | 6718 hCoV-19/England/CAMC-BBCD10/2020\|EPI_ISL_679276\|20 | ·················· | ·A················ | ·················· | ·················· | ·················· | ··A··············· | |
| U | 6719 hCoV-19/England/CAMC-BBCD2F/2020\|EPI_ISL_679277\|20 | ·················· | ·················· | ·················· | ·················· | ·················· | ·················· | |
| U | 6720 hCoV-19/England/CAMC-BBCD5C/2020\|EPI_ISL_679280\|20 | ·················· | ·················· | ·················· | ·················· | ·················· | ·················· | |
| U | 6721 hCoV-19/England/CAMC-BBCD6B/2020\|EPI_ISL_679281\|20 | ·················· | ·················· | ·················· | ·················· | ·················· | ·················· | |
| U | 6722 hCoV-19/England/CAMC-BBCDE3/2020\|EPI_ISL_679284\|20 | ·················· | ·················· | ·················· | ·················· | ·················· | ·················· | |
| U | 6723 hCoV-19/England/CAMC-BBCE1D/2020\|EPI_ISL_679287\|20 | ·················· | ·················· | ·················· | ·················· | ·················· | ·················· | |
| U | 6724 hCoV-19/England/CAMC-BBCE4A/2020\|EPI_ISL_679289\|20 | ·················· | ·················· | ·················· | ·················· | ·················· | ·················· | |
| U | 6725 hCoV-19/England/CAMC-BBCE77/2020\|EPI_ISL_679291\|20 | ·················· | ·················· | ·················· | ·················· | ·················· | ·················· | |
| U | 6726 hCoV-19/England/CAMC-BBCEE0/2020\|EPI_ISL_679294\|20 | ·················· | ·················· | ·················· | ·················· | ·················· | ·················· | |
| U | 6727 hCoV-19/England/CAMC-BBCF74/2020\|EPI_ISL_679296\|20 | ·················· | ·················· | ·················· | ·················· | ·················· | ·················· | |
| U | 6728 hCoV-19/England/CAMC-BBD15F/2020\|EPI_ISL_679297\|20 | ·················· | ·················· | ·················· | ·················· | ·················· | ·················· | |
| U | 6729 hCoV-19/England/CAMC-BBD1D7/2020\|EPI_ISL_679303\|20 | ·················· | ·················· | ·················· | ·················· | ·················· | ·················· | |
| U | 6730 hCoV-19/England/CAMC-BBD1F5/2020\|EPI_ISL_679305\|20 | ·················· | ·················· | ·················· | ·················· | ·················· | ·················· | |
| U | 6731 hCoV-19/England/CAMC-BBD22F/2020\|EPI_ISL_679307\|20 | ·················· | ·················· | ·················· | ·················· | ·················· | ·················· | |
| U | 6732 hCoV-19/England/CAMC-BBD24D/2020\|EPI_ISL_679308\|20 | ·················· | ·················· | ·················· | ·················· | ·················· | ·················· | |
| U | 6733 hCoV-19/England/CAMC-BBD26B/2020\|EPI_ISL_679309\|20 | ·················· | ·················· | ·················· | ·················· | ·················· | ·················· | |
| U | 6734 hCoV-19/England/CAMC-BBD2B6/2020\|EPI_ISL_679310\|20 | ·················· | ·················· | ·················· | ·················· | ·················· | ·················· | |
| U | 6735 hCoV-19/England/CAMC-BBD2C5/2020\|EPI_ISL_679311\|20 | ·················· | ·················· | ·················· | ·················· | ·················· | ·················· | |
| U | 6736 hCoV-19/England/CAMC-BBD4CF/2020\|EPI_ISL_679314\|20 | ·················· | ·················· | ·················· | ·················· | ·················· | ·················· | |
| U | 6737 hCoV-19/England/EXET-139B93/2020\|EPI_ISL_679327\|20 | ·················· | ·················· | ·················· | ·················· | ·················· | ·················· | |
| U | 6738 hCoV-19/England/EXET-139C90/2020\|EPI_ISL_679332\|20 | ·················· | ·················· | ·················· | ·················· | ·················· | ·················· | |
| U | 6739 hCoV-19/England/EXET-139D15/2020\|EPI_ISL_679337\|20 | ·················· | ·················· | ·················· | ·················· | ·················· | ·················· | |
| U | 6740 hCoV-19/Northern Ireland/NIRE-232E81/2020\|EPI_ISL_ | ·················· | ·················· | ·················· | ·················· | ·················· | ·················· | |
| U | 6741 hCoV-19/Northern Ireland/NIRE-232E90/2020\|EPI_ISL_ | ·················· | ·················· | ·················· | ·················· | ·················· | ·················· | |
| U | 6742 hCoV-19/Northern Ireland/NIRE-232EBE/2020\|EPI_ISL_ | ·················· | ·················· | ·················· | ·················· | ·················· | ·················· | |
| U | 6745 hCoV-19/Northern Ireland/NIRE-232FAC/2020\|EPI_ISL_ | ·················· | ·················· | ·················· | ·················· | ·················· | ·················· | |
| U | 6754 hCoV-19/Northern Ireland/NIRE-233099/2020\|EPI_ISL_ | ·················· | ·················· | ·················· | ·················· | ·················· | ·················· | |
| U | 6755 hCoV-19/Northern Ireland/NIRE-2330A8/2020\|EPI_ISL_ | ·················· | ·················· | ·················· | ·················· | ·················· | ·················· | |
| U | 6759 hCoV-19/Northern Ireland/NIRE-2331C3/2020\|EPI_ISL_ | ·················· | ·················· | ·················· | ·················· | NNN··············· | ·················· | 1 |
| U | 6761 hCoV-19/Northern Ireland/NIRE-23321B/2020\|EPI_ISL_ | ·················· | ·················· | ·················· | ·················· | NNN··············· | ·················· | 1 |
| U | 6767 hCoV-19/Northern Ireland/NIRE-233293/2020\|EPI_ISL_ | ·················· | ·················· | ·················· | ·················· | ·················· | ·················· | |
| U | 6773 hCoV-19/Northern Ireland/NIRE-233336/2020\|EPI_ISL_ | ·················· | ·················· | ·················· | ·················· | ·················· | ·················· | |
| U | 6777 hCoV-19/Northern Ireland/NIRE-233451/2020\|EPI_ISL_ | ·················· | ·················· | ·················· | ·················· | NNN··············· | ·················· | 1 |
| U | 6778 hCoV-19/Northern Ireland/NIRE-2334CA/2020\|EPI_ISL_ | ·················· | ·A················ | ·················· | ·················· | ·················· | ·················· | |
| U | 6779 hCoV-19/Northern Ireland/NIRE-2334E8/2020\|EPI_ISL_ | ·················· | ·················· | ·················· | ·················· | ·················· | ·················· | |
| U | 6780 hCoV-19/Northern Ireland/NIRE-233512/2020\|EPI_ISL_ | ·················· | ·················· | ·················· | ·················· | ·················· | ·················· | |
| U | 6781 hCoV-19/Northern Ireland/NIRE-2336C4/2020\|EPI_ISL_ | ·················· | ·················· | ·················· | ·················· | ·················· | ·················· | |
| U | 6782 hCoV-19/Northern Ireland/NIRE-2336E2/2020\|EPI_ISL_ | ·················· | ·················· | ·················· | ·················· | ·················· | ·················· | |
| U | 6783 hCoV-19/Northern Ireland/NIRE-23370D/2020\|EPI_ISL_ | ·················· | ·················· | ·················· | ·················· | ·················· | ·················· | |
| U | 6784 hCoV-19/Northern Ireland/NIRE-233749/2020\|EPI_ISL_ | ·················· | ·················· | ·················· | ·················· | ·················· | ·················· | |
| U | 6785 hCoV-19/Northern Ireland/NIRE-233767/2020\|EPI_ISL_ | ·················· | ·················· | ·················· | ·················· | ·················· | ·················· | |
| U | 6786 hCoV-19/Northern Ireland/NIRE-233776/2020\|EPI_ISL_ | ·················· | ·················· | ·················· | ·················· | ·················· | ·················· | |
| U | 6787 hCoV-19/Northern Ireland/NIRE-233785/2020\|EPI_ISL_ | ·················· | ·················· | ·················· | ·················· | ·················· | ·················· | |
| U | 6788 hCoV-19/Northern Ireland/NIRE-2337B2/2020\|EPI_ISL_ | ·················· | ·················· | ·················· | ·················· | ·················· | ·················· | |
| U | 6789 hCoV-19/Northern Ireland/NIRE-233855/2020\|EPI_ISL_ | ·················· | ·················· | ·················· | ·················· | ·················· | ·················· | |
| U | 6790 hCoV-19/Northern Ireland/NIRE-233907/2020\|EPI_ISL_ | ·················· | ·················· | ·················· | ·················· | ·················· | ·················· | |
| U | 6791 hCoV-19/Northern Ireland/NIRE-233952/2020\|EPI_ISL_ | ·················· | ·················· | ·················· | ·················· | ·················· | ·················· | |
| U | 6793 hCoV-19/England/MILK-BAB1EB/2020\|EPI_ISL_681562\|20 | ·················· | ·················· | ·················· | ·················· | ·················· | ·················· | |

| | | | | | | | |
|---|---|---|---|---|---|---|---|
| U | 6794 hCoV-19/England/MILK-BA9B2B/2020\|EPI_ISL_681566\|20 | .................... | .................... | .................... | .................... | .................... | .................... |
| U | 6795 hCoV-19/England/MILK-BACD31/2020\|EPI_ISL_681569\|20 | .................... | .................... | .................... | .................... | .................... | .................... |
| U | 6796 hCoV-19/England/MILK-BAB576/2020\|EPI_ISL_681570\|20 | .................... | .................... | .................... | .................... | .................... | .................... |
| U | 6797 hCoV-19/England/MILK-BAB655/2020\|EPI_ISL_681571\|20 | .................... | .................... | .................... | .................... | .................... | .................... |
| U | 6798 hCoV-19/England/MILK-BAB4A6/2020\|EPI_ISL_681572\|20 | .................... | .................... | .................... | .................... | .................... | .................... |
| U | 6799 hCoV-19/England/MILK-BAAC45/2020\|EPI_ISL_681573\|20 | .................... | .................... | .................... | .................... | .................... | .................... |
| U | 6800 hCoV-19/England/MILK-BACD8C/2020\|EPI_ISL_681574\|20 | .................... | .................... | .................... | .................... | .................... | .................... |
| U | 6801 hCoV-19/England/MILK-BAA7EA/2020\|EPI_ISL_681575\|20 | .................... | .................... | .................... | .................... | .................... | .................... |
| U | 6802 hCoV-19/England/MILK-BAB497/2020\|EPI_ISL_681576\|20 | .................... | .................... | .................... | .................... | .................... | .................... |
| U | 6803 hCoV-19/England/MILK-BAAE4F/2020\|EPI_ISL_681577\|20 | .................... | .................... | .................... | .................... | .................... | .................... |
| U | 6804 hCoV-19/England/MILK-BA9B67/2020\|EPI_ISL_681578\|20 | .................... | .................... | .................... | .................... | .................... | .................... |
| U | 6805 hCoV-19/England/MILK-BAAAB4/2020\|EPI_ISL_681579\|20 | .................... | .................... | .................... | .................... | .................... | .................... |
| U | 6806 hCoV-19/England/MILK-BA9EE6/2020\|EPI_ISL_681580\|20 | .................... | .................... | .................... | .................... | .................... | .................... |
| U | 6807 hCoV-19/England/MILK-BABBB0/2020\|EPI_ISL_681582\|20 | .................... | .................... | .................... | .................... | .................... | .................... |
| U | 6808 hCoV-19/England/MILK-BAAFE2/2020\|EPI_ISL_681583\|20 | .................... | .................... | .................... | .................... | .................... | .................... |
| U | 6809 hCoV-19/England/MILK-BAB52B/2020\|EPI_ISL_681584\|20 | .................... | .................... | .................... | .................... | .................... | .................... |
| U | 6810 hCoV-19/England/MILK-BAAB2A/2020\|EPI_ISL_681586\|20 | .................... | .................... | .................... | .................... | .................... | .................... |
| U | 6811 hCoV-19/England/MILK-BAA86F/2020\|EPI_ISL_681587\|20 | .................... | .................... | .................... | .................... | .................... | .................... |
| U | 6812 hCoV-19/England/MILK-BA9ED7/2020\|EPI_ISL_681589\|20 | .................... | .................... | .................... | .................... | .................... | .................... |
| U | 6813 hCoV-19/England/MILK-BA9CFB/2020\|EPI_ISL_681590\|20 | .................... | .................... | .................... | .................... | .................... | .................... |
| U | 6814 hCoV-19/England/MILK-BAA8E7/2020\|EPI_ISL_681591\|20 | .................... | .................... | .................... | .................... | .................... | .................... |
| U | 6815 hCoV-19/England/MILK-BA9BB2/2020\|EPI_ISL_681592\|20 | .................... | .................... | .................... | .................... | .................... | .................... |
| U | 6816 hCoV-19/England/MILK-BAC6EB/2020\|EPI_ISL_681593\|20 | .................... | .................... | .................... | .................... | .................... | .................... |
| U | 6817 hCoV-19/England/MILK-BAABC0/2020\|EPI_ISL_681594\|20 | .................... | .A.................. | .................... | .................... | .................... | ..A................. |
| U | 6818 hCoV-19/England/MILK-BAA9F3/2020\|EPI_ISL_681597\|20 | .................... | .................... | .................... | .................... | .................... | .................... |
| U | 6819 hCoV-19/England/MILK-BABE3F/2020\|EPI_ISL_681598\|20 | .................... | .................... | .................... | .................... | .................... | .................... |
| U | 6820 hCoV-19/England/MILK-BABE8A/2020\|EPI_ISL_681600\|20 | .................... | .................... | .................... | .................... | .................... | .................... |
| U | 6821 hCoV-19/England/MILK-BAA98A/2020\|EPI_ISL_681601\|20 | .................... | .................... | .................... | .................... | .................... | .................... |
| U | 6822 hCoV-19/England/MILK-BAB51C/2020\|EPI_ISL_681602\|20 | .................... | .................... | .................... | .................... | .................... | .................... |
| U | 6823 hCoV-19/England/MILK-BAB488/2020\|EPI_ISL_681603\|20 | .................... | .................... | .................... | .................... | .................... | .................... |
| U | 6824 hCoV-19/England/MILK-BAB6CE/2020\|EPI_ISL_681604\|20 | .................... | .................... | .................... | .................... | .................... | .................... |
| U | 6825 hCoV-19/England/MILK-BAB45B/2020\|EPI_ISL_681605\|20 | .................... | .................... | .................... | .................... | .................... | .................... |
| U | 6826 hCoV-19/England/MILK-BAB190/2020\|EPI_ISL_681607\|20 | .................... | .................... | .................... | .................... | .................... | .................... |
| U | 6827 hCoV-19/England/MILK-BA9F2F/2020\|EPI_ISL_681608\|20 | .................... | .................... | .................... | .................... | .................... | .................... |
| U | 6828 hCoV-19/England/MILK-BAAA4B/2020\|EPI_ISL_681610\|20 | .................... | .................... | .................... | .................... | .................... | .................... |
| U | 6829 hCoV-19/England/MILK-BAB7CB/2020\|EPI_ISL_681611\|20 | .................... | .................... | .................... | .................... | .................... | .................... |
| U | 6830 hCoV-19/England/MILK-BAC672/2020\|EPI_ISL_681613\|20 | .................... | .A.................. | .................... | .................... | .................... | ..A................. |
| U | 6831 hCoV-19/England/MILK-BAAFA6/2020\|EPI_ISL_681614\|20 | .................... | .................... | .................... | .................... | .....T.............. | .................... |
| U | 6832 hCoV-19/England/MILK-BAABFD/2020\|EPI_ISL_681615\|20 | .................... | .................... | .................... | .................... | .................... | .................... |
| U | 6833 hCoV-19/England/MILK-BAC6DC/2020\|EPI_ISL_681616\|20 | .................... | .................... | .................... | .................... | .................... | .................... |
| U | 6834 hCoV-19/England/MILK-BAAA69/2020\|EPI_ISL_681617\|20 | .................... | .................... | .................... | .................... | .................... | .................... |
| U | 6835 hCoV-19/England/MILK-BA9B0D/2020\|EPI_ISL_681618\|20 | .................... | .................... | .................... | .................... | .................... | .................... |
| U | 6836 hCoV-19/England/MILK-BAA02B/2020\|EPI_ISL_681620\|20 | .................... | .................... | .................... | .................... | .................... | .................... |
| U | 6837 hCoV-19/England/MILK-BA9F7A/2020\|EPI_ISL_681621\|20 | .................... | .................... | .................... | .................... | .................... | .................... |
| U | 6838 hCoV-19/England/MILK-BAB4D3/2020\|EPI_ISL_681622\|20 | .................... | .................... | .................... | .................... | .................... | .................... |
| U | 6839 hCoV-19/England/MILK-BAA771/2020\|EPI_ISL_681623\|20 | .................... | .................... | .................... | .................... | .................... | .................... |
| U | 6840 hCoV-19/England/MILK-BAB224/2020\|EPI_ISL_681624\|20 | .................... | .A.................. | .................... | .................... | .................... | ..A................. |
| U | 6841 hCoV-19/England/MILK-BACD5F/2020\|EPI_ISL_681625\|20 | .................... | .................... | .................... | .................... | .................... | .................... |
| U | 6842 hCoV-19/England/MILK-BAA559/2020\|EPI_ISL_681626\|20 | .................... | .................... | .................... | .................... | .................... | .................... |
| U | 6843 hCoV-19/England/MILK-BAB29D/2020\|EPI_ISL_681627\|20 | .................... | .................... | .................... | .................... | .................... | .................... |
| U | 6844 hCoV-19/England/MILK-BAA191/2020\|EPI_ISL_681628\|20 | .................... | .................... | .................... | .................... | .................... | .................... |
| U | 6845 hCoV-19/England/MILK-BAAA5A/2020\|EPI_ISL_681630\|20 | .................... | .................... | .................... | .................... | .................... | .................... |
| U | 6846 hCoV-19/England/MILK-BAB6A0/2020\|EPI_ISL_681631\|20 | .................... | .................... | .................... | .................... | .................... | .................... |
| U | 6847 hCoV-19/England/MILK-BAB567/2020\|EPI_ISL_681632\|20 | .................... | .................... | .................... | .................... | .................... | .................... |
| U | 6848 hCoV-19/England/MILK-BAA9A8/2020\|EPI_ISL_681633\|20 | .................... | .................... | .................... | .................... | .................... | .................... |
| U | 6849 hCoV-19/England/MILK-BACD9B/2020\|EPI_ISL_681635\|20 | .................... | .................... | .................... | .................... | .................... | .................... |
| U | 6850 hCoV-19/England/MILK-BA9A6A/2020\|EPI_ISL_681636\|20 | .................... | .................... | .................... | .................... | .................... | .................... |
| U | 6851 hCoV-19/England/MILK-BAAFB5/2020\|EPI_ISL_681638\|20 | .................... | .................... | .................... | .................... | .................... | .................... |
| U | 6852 hCoV-19/England/MILK-BAC44B/2020\|EPI_ISL_681639\|20 | .................... | .................... | .................... | .................... | .................... | .................... |
| U | 6853 hCoV-19/England/MILK-BAC636/2020\|EPI_ISL_681640\|20 | .................... | .................... | .................... | .................... | .................... | .................... |
| U | 6854 hCoV-19/England/MILK-BA9BA3/2020\|EPI_ISL_681641\|20 | .................... | .................... | .................... | .................... | .................... | .................... |
| U | 6855 hCoV-19/England/MILK-BA9F89/2020\|EPI_ISL_681642\|20 | .................... | .................... | .................... | .................... | .................... | .................... |
| U | 6856 hCoV-19/England/MILK-BABE11/2020\|EPI_ISL_681644\|20 | .................... | .................... | .................... | .................... | .................... | .................... |
| U | 6857 hCoV-19/England/MILK-BAA999/2020\|EPI_ISL_681646\|20 | .................... | .................... | .................... | .................... | .................... | .................... |
| U | 6858 hCoV-19/England/MILK-BA9EB9/2020\|EPI_ISL_681648\|20 | .................... | .................... | .................... | .................... | .................... | .................... |
| U | 6859 hCoV-19/England/MILK-BAB27F/2020\|EPI_ISL_681650\|20 | .................... | .................... | .................... | .................... | .................... | .................... |
| U | 6860 hCoV-19/England/MILK-BA9B49/2020\|EPI_ISL_681651\|20 | .................... | .................... | .................... | .................... | .................... | .................... |
| U | 6861 hCoV-19/England/MILK-BAB43D/2020\|EPI_ISL_681652\|20 | .................... | .................... | .................... | .................... | .................... | .................... |
| U | 6862 hCoV-19/England/MILK-BAB619/2020\|EPI_ISL_681654\|20 | .................... | .................... | .................... | .................... | .................... | .................... |
| U | 6863 hCoV-19/England/MILK-BAAA78/2020\|EPI_ISL_681655\|20 | .................... | .................... | .................... | .................... | .................... | .................... |
| U | 6864 hCoV-19/England/CAMC-B08FF1/2020\|EPI_ISL_681656\|20 | .................... | .................... | .................... | .................... | .................... | .................... |
| U | 6865 hCoV-19/England/CAMC-B9C596/2020\|EPI_ISL_681659\|20 | .................... | .................... | .................... | .................... | .................... | .................... |
| U | 6866 hCoV-19/Denmark/DCGC-12712/2020\|EPI_ISL_682893\|202 | .................... | .................... | .................... | .................... | .................... | .................... |
| U | 6867 hCoV-19/Denmark/DCGC-12713/2020\|EPI_ISL_682894\|202 | .................... | .................... | .................... | .................... | .................... | .................... |
| U | 6868 hCoV-19/Denmark/DCGC-12714/2020\|EPI_ISL_682895\|202 | .................... | .................... | .................... | .................... | .................... | .................... |
| U | 6869 hCoV-19/Denmark/DCGC-12716/2020\|EPI_ISL_682897\|202 | .................... | .................... | .................... | .................... | .................... | .................... |
| U | 6870 hCoV-19/Denmark/DCGC-12717/2020\|EPI_ISL_682898\|202 | .................... | .................... | .................... | .................... | .................... | .................... |
| U | 6871 hCoV-19/Denmark/DCGC-12718/2020\|EPI_ISL_682899\|202 | .................... | .................... | .................... | .................... | .................... | .................... |
| U | 6872 hCoV-19/Denmark/DCGC-12752/2020\|EPI_ISL_682933\|202 | .................... | .................... | .................... | .................... | .................... | .................... |
| U | 6873 hCoV-19/Denmark/DCGC-12753/2020\|EPI_ISL_682934\|202 | .................... | .................... | .................... | .................... | .................... | .................... |

| | | | | | | | |
|---|---|---|---|---|---|---|---|
| U | 6874 hCoV-19/Denmark/DCGC-12754/2020\|EPI_ISL_682935\|202 | ···················· | ···················· | ····················· | ···················· | ···················· | ···················· |
| U | 6875 hCoV-19/Denmark/DCGC-12755/2020\|EPI_ISL_682936\|202 | ···················· | ···················· | ····················· | ···················· | ···················· | ···················· |
| U | 6876 hCoV-19/Denmark/DCGC-12758/2020\|EPI_ISL_682939\|202 | ···················· | ···················· | ····················· | ···················· | ···················· | ···················· |
| U | 6877 hCoV-19/Denmark/DCGC-12759/2020\|EPI_ISL_682940\|202 | ···················· | ···················· | ····················· | ···················· | ···················· | ···················· |
| U | 6878 hCoV-19/Denmark/DCGC-12760/2020\|EPI_ISL_682941\|202 | ···················· | ·A·················· | ····················· | ···················· | ···················· | ···················· |
| U | 6879 hCoV-19/Denmark/DCGC-12761/2020\|EPI_ISL_682942\|202 | ···················· | ···················· | ····················· | ···················· | ···················· | ···················· |
| U | 6880 hCoV-19/Denmark/DCGC-12763/2020\|EPI_ISL_682944\|202 | ···················· | ···················· | ····················· | ···················· | ···················· | ···················· |
| U | 6881 hCoV-19/Denmark/DCGC-12764/2020\|EPI_ISL_682945\|202 | ···················· | ···················· | ····················· | ···················· | ···················· | ···················· |
| U | 6882 hCoV-19/Denmark/DCGC-12765/2020\|EPI_ISL_682946\|202 | ···················· | ···················· | ····················· | ···················· | ···················· | ···················· |
| U | 6883 hCoV-19/Denmark/DCGC-12766/2020\|EPI_ISL_682947\|202 | ···················· | ···················· | ····················· | ···················· | ···················· | ···················· |
| U | 6884 hCoV-19/Denmark/DCGC-12767/2020\|EPI_ISL_682948\|202 | ···················· | ···················· | ····················· | ···················· | ···················· | ···················· |
| U | 6885 hCoV-19/Denmark/DCGC-12770/2020\|EPI_ISL_682951\|202 | ···················· | ···················· | ····················· | ···················· | ···················· | ···················· |
| U | 6886 hCoV-19/Denmark/DCGC-12771/2020\|EPI_ISL_682952\|202 | ···················· | ···················· | ····················· | ···················· | ···················· | ···················· |
| U | 6887 hCoV-19/Denmark/DCGC-12772/2020\|EPI_ISL_682953\|202 | ···················· | ···················· | ····················· | ···················· | ···················· | ···················· |
| U | 6888 hCoV-19/Denmark/DCGC-12773/2020\|EPI_ISL_682954\|202 | ···················· | ···················· | ····················· | ···················· | ···················· | ···················· |
| U | 6889 hCoV-19/Denmark/DCGC-12777/2020\|EPI_ISL_682958\|202 | ···················· | ···················· | ····················· | ···················· | ···················· | ···················· |
| U | 6890 hCoV-19/Denmark/DCGC-12780/2020\|EPI_ISL_682961\|202 | ···················· | ···················· | ····················· | ···················· | ···················· | ···················· |
| U | 6891 hCoV-19/Denmark/DCGC-12781/2020\|EPI_ISL_682962\|202 | ···················· | ···················· | ····················· | ···················· | ···················· | ···················· |
| U | 6892 hCoV-19/Denmark/DCGC-12782/2020\|EPI_ISL_682963\|202 | ···················· | ···················· | ····················· | ···················· | ···················· | ···················· |
| U | 6893 hCoV-19/Denmark/DCGC-12783/2020\|EPI_ISL_682964\|202 | ···················· | ···················· | ····················· | ···················· | ···················· | ···················· |
| U | 6894 hCoV-19/Denmark/DCGC-12784/2020\|EPI_ISL_682965\|202 | ···················· | ···················· | ····················· | ···················· | ···················· | ···················· |
| U | 6895 hCoV-19/Denmark/DCGC-12785/2020\|EPI_ISL_682966\|202 | ···················· | ···················· | ····················· | ···················· | ···················· | ···················· |
| U | 6896 hCoV-19/Denmark/DCGC-12786/2020\|EPI_ISL_682967\|202 | ···················· | ···················· | ····················· | ···················· | ···················· | ···················· |
| U | 6897 hCoV-19/Denmark/DCGC-12787/2020\|EPI_ISL_682968\|202 | ···················· | ···················· | ····················· | ···················· | ···················· | ···················· |
| U | 6898 hCoV-19/Denmark/DCGC-12788/2020\|EPI_ISL_682969\|202 | ···················· | ···················· | ····················· | ···················· | ···················· | ···················· |
| U | 6899 hCoV-19/Denmark/DCGC-12789/2020\|EPI_ISL_682970\|202 | ···················· | ····A··············· | ····················· | ···················· | ···················· | ···················· |
| U | 6900 hCoV-19/Denmark/DCGC-12790/2020\|EPI_ISL_682971\|202 | ···················· | ···················· | ····················· | ···················· | ···················· | ···················· |
| U | 6901 hCoV-19/Denmark/DCGC-12791/2020\|EPI_ISL_682972\|202 | ···················· | ···················· | ····················· | ···················· | ···················· | ···················· |
| U | 6902 hCoV-19/Denmark/DCGC-12792/2020\|EPI_ISL_682973\|202 | ···················· | ···················· | ····················· | ···················· | ···················· | ···················· |
| U | 6903 hCoV-19/Denmark/DCGC-12793/2020\|EPI_ISL_682974\|202 | ···················· | ···················· | ····················· | ···················· | ···················· | ···················· |
| U | 6904 hCoV-19/Denmark/DCGC-12794/2020\|EPI_ISL_682975\|202 | ···················· | ···················· | ····················· | ···················· | ···················· | ···················· |
| U | 6905 hCoV-19/Denmark/DCGC-12795/2020\|EPI_ISL_682976\|202 | ···················· | ···················· | ····················· | ···················· | ···················· | ···················· |
| U | 6906 hCoV-19/Denmark/DCGC-12798/2020\|EPI_ISL_682979\|202 | ···················· | ···················· | ····················· | ···················· | ···················· | ···················· |
| U | 6907 hCoV-19/Denmark/DCGC-12800/2020\|EPI_ISL_682981\|202 | ···················· | ···················· | ····················· | ···················· | ···················· | ···················· |
| U | 6908 hCoV-19/Denmark/DCGC-12803/2020\|EPI_ISL_682984\|202 | ···················· | ···················· | ····················· | ···················· | ···················· | ···················· |
| U | 6909 hCoV-19/Denmark/DCGC-12804/2020\|EPI_ISL_682985\|202 | ···················· | ···················· | ····················· | ···················· | ···················· | ···················· |
| U | 6910 hCoV-19/Denmark/DCGC-12805/2020\|EPI_ISL_682986\|202 | ···················· | ···················· | ····················· | ···················· | ···················· | ···················· |
| U | 6911 hCoV-19/Denmark/DCGC-12806/2020\|EPI_ISL_682987\|202 | ···················· | ···················· | ····················· | ···················· | ···················· | ···················· |
| U | 6912 hCoV-19/Denmark/DCGC-12807/2020\|EPI_ISL_682988\|202 | ···················· | ···················· | ····················· | ···················· | ···················· | ···················· |
| U | 6913 hCoV-19/England/CAMC-B096FB/2020\|EPI_ISL_673026\|20 | ···················· | ···················· | ····················· | ···················· | ···················· | ···················· |
| U | 6914 hCoV-19/England/CAMC-B09136/2020\|EPI_ISL_673027\|20 | ···················· | ···················· | ····················· | ···················· | ······T············· | ···················· |
| U | 6915 hCoV-19/England/CAMC-B0A108/2020\|EPI_ISL_673028\|20 | ···················· | ···················· | ····················· | ···················· | ···················· | ···················· |
| U | 6916 hCoV-19/England/CAMC-B0944C/2020\|EPI_ISL_673029\|20 | ···················· | ···················· | ····················· | ···················· | ···················· | ···················· |
| U | 6917 hCoV-19/England/CAMC-B09215/2020\|EPI_ISL_673030\|20 | ···················· | ···················· | ····················· | ···················· | ···················· | ···················· |
| U | 6918 hCoV-19/England/CAMC-B0A056/2020\|EPI_ISL_673031\|20 | ···················· | ···················· | ····················· | ···················· | ···················· | ···················· |
| U | 6919 hCoV-19/England/CAMC-B0A40F/2020\|EPI_ISL_673032\|20 | ···················· | ···················· | ····················· | ···················· | ···················· | ···················· |
| U | 6920 hCoV-19/England/CAMC-B08F79/2020\|EPI_ISL_673033\|20 | ···················· | ···················· | ····················· | ···················· | ····T··············· | ···················· |
| U | 6921 hCoV-19/England/CAMC-B099E3/2020\|EPI_ISL_673035\|20 | ···················· | ···················· | ····················· | ···················· | ···················· | ···················· |
| U | 6922 hCoV-19/England/CAMC-B09655/2020\|EPI_ISL_673036\|20 | ···················· | ···················· | ····················· | ···················· | ···················· | ···················· |
| U | 6923 hCoV-19/England/CAMC-B08F88/2020\|EPI_ISL_673037\|20 | ···················· | ···················· | ····················· | ···················· | ···················· | ···················· |
| U | 6924 hCoV-19/England/CAMC-B09D9C/2020\|EPI_ISL_673038\|20 | ···················· | ···················· | ····················· | ···················· | ······T············· | ···················· |
| U | 6925 hCoV-19/England/CAMC-B09E5D/2020\|EPI_ISL_673039\|20 | ···················· | ···················· | ····················· | ···················· | ······T············· | ···················· |
| U | 6926 hCoV-19/England/CAMC-B09109/2020\|EPI_ISL_673040\|20 | ···················· | ···················· | ····················· | ···················· | ···················· | ···················· |
| U | 6927 hCoV-19/England/CAMC-B09743/2020\|EPI_ISL_673041\|20 | ···················· | ···················· | ····················· | ···················· | ···················· | ···················· |
| U | 6928 hCoV-19/England/CAMC-B0986E/2020\|EPI_ISL_673042\|20 | ···················· | ···················· | ····················· | ···················· | ···················· | ···················· |
| U | 6929 hCoV-19/England/CAMC-B08D24/2020\|EPI_ISL_673043\|20 | ···················· | ···················· | ····················· | ···················· | ···················· | ···················· |
| U | 6930 hCoV-19/England/CAMC-B098B9/2020\|EPI_ISL_673044\|20 | ···················· | ···················· | ····················· | ···················· | ···················· | ···················· |
| U | 6931 hCoV-19/England/CAMC-B08B93/2020\|EPI_ISL_673046\|20 | ···················· | ···················· | ····················· | ···················· | ···················· | ···················· |
| U | 6932 hCoV-19/England/CAMC-B09FB4/2020\|EPI_ISL_673047\|20 | ···················· | ···················· | ····················· | ···················· | ···················· | ···················· |
| U | 6933 hCoV-19/England/CAMC-B0A4B4/2020\|EPI_ISL_673048\|20 | ···················· | ···················· | ····················· | ···················· | ···················· | ···················· |
| U | 6934 hCoV-19/England/CAMC-B0A5B1/2020\|EPI_ISL_673050\|20 | ···················· | ···················· | ····················· | ···················· | ···················· | ···················· |
| U | 6935 hCoV-19/England/CAMC-B08F97/2020\|EPI_ISL_673051\|20 | ···················· | ···················· | ····················· | ···················· | ···················· | ···················· |
| U | 6936 hCoV-19/England/CAMC-B0A19F/2020\|EPI_ISL_673052\|20 | ···················· | ···················· | ····················· | ···················· | ···················· | ···················· |
| U | 6937 hCoV-19/England/CAMC-B08EB8/2020\|EPI_ISL_673053\|20 | ···················· | ···················· | ····················· | ···················· | ······T············· | ···················· |
| U | 6938 hCoV-19/England/CAMC-B09558/2020\|EPI_ISL_673054\|20 | ···················· | ···················· | ····················· | ···················· | ···················· | ···················· |
| U | 6939 hCoV-19/England/CAMC-B0A092/2020\|EPI_ISL_673055\|20 | ···················· | ···················· | ····················· | ···················· | ···················· | ···················· |
| U | 6940 hCoV-19/England/CAMC-B0A5FD/2020\|EPI_ISL_673056\|20 | ···················· | ···················· | ····················· | ···················· | ···················· | ···················· |
| U | 6941 hCoV-19/England/CAMC-B099B6/2020\|EPI_ISL_673057\|20 | ···················· | ···················· | ····················· | ···················· | ···················· | ···················· |
| U | 6942 hCoV-19/England/CAMC-B08B84/2020\|EPI_ISL_673058\|20 | ···················· | ···················· | ····················· | ···················· | ···················· | ···················· |
| U | 6943 hCoV-19/England/CAMC-B09B83/2020\|EPI_ISL_673059\|20 | ···················· | ···················· | ····················· | ···················· | ···················· | ···················· |
| U | 6944 hCoV-19/England/CAMC-B0A44B/2020\|EPI_ISL_673062\|20 | ···················· | ···················· | ····················· | ···················· | ···················· | ···················· |
| U | 6945 hCoV-19/England/CAMC-B0939A/2020\|EPI_ISL_673063\|20 | ···················· | ···················· | ····················· | ···················· | ···················· | ···················· |
| U | 6946 hCoV-19/England/CAMC-B09F4B/2020\|EPI_ISL_673064\|20 | ···················· | ···················· | ····················· | ···················· | ···················· | ···················· |
| U | 6947 hCoV-19/England/CAMC-B093D6/2020\|EPI_ISL_673065\|20 | ···················· | ···················· | ····················· | ···················· | ···················· | ···················· |
| U | 6948 hCoV-19/England/CAMC-B09C35/2020\|EPI_ISL_673066\|20 | ···················· | ···················· | ····················· | ···················· | ···················· | ···················· |
| U | 6949 hCoV-19/England/CAMC-B0A496/2020\|EPI_ISL_673067\|20 | ···················· | ···················· | ····················· | ···················· | ···················· | ···················· |
| U | 6950 hCoV-19/England/CAMC-B09BB0/2020\|EPI_ISL_673068\|20 | ···················· | ···················· | ····················· | ···················· | ···················· | ···················· |
| U | 6951 hCoV-19/England/CAMC-B0A41E/2020\|EPI_ISL_673069\|20 | ···················· | ···················· | ····················· | ···················· | ···················· | ···················· |
| U | 6952 hCoV-19/England/CAMC-B09084/2020\|EPI_ISL_673070\|20 | ···················· | ···················· | ····················· | ···················· | ···················· | ···················· |
| U | 6953 hCoV-19/England/CAMC-B09813/2020\|EPI_ISL_673072\|20 | ···················· | ···················· | ····················· | ···················· | ···················· | ···················· |

| | | | | | | | | |
|---|---|---|---|---|---|---|---|---|
| U | 6954 | hCoV-19/England/CAMC-B08E9A/2020\|EPI_ISL_673073\|20 | ···················· | ···················· | ····················· | ···················· | ···················· | ···················· |
| U | 6955 | hCoV-19/England/CAMC-B09E99/2020\|EPI_ISL_673074\|20 | ···················· | ···················· | ····················· | ···················· | ···················· | ···················· |
| U | 6956 | hCoV-19/England/CAMC-B08F5B/2020\|EPI_ISL_673076\|20 | ···················· | ···················· | ····················· | ···················· | ···················· | ···················· |
| U | 6957 | hCoV-19/England/CAMC-B0A5C0/2020\|EPI_ISL_673077\|20 | ···················· | ···················· | ····················· | ···················· | ···················· | ···················· |
| U | 6958 | hCoV-19/England/CAMC-B0A566/2020\|EPI_ISL_673079\|20 | ···················· | ···················· | ····················· | ···················· | ···················· | ···················· |
| U | 6959 | hCoV-19/England/CAMC-B08EE5/2020\|EPI_ISL_673081\|20 | ···················· | ···················· | ····················· | ···················· | ···················· | ···················· |
| U | 6960 | hCoV-19/England/CAMC-B0A609/2020\|EPI_ISL_673082\|20 | ···················· | ···················· | ····················· | ···················· | ···················· | ···················· |
| U | 6961 | hCoV-19/England/CAMC-B09D50/2020\|EPI_ISL_673083\|20 | ···················· | ···················· | ····················· | ···················· | ···················· | ···················· |
| U | 6962 | hCoV-19/England/CAMC-B09DD8/2020\|EPI_ISL_673084\|20 | ···················· | ···················· | ····················· | ···················· | ···················· | ···················· |
| U | 6963 | hCoV-19/England/CAMC-B0A627/2020\|EPI_ISL_673085\|20 | ···················· | ···················· | ····················· | ···················· | ···················· | ···················· |
| U | 6964 | hCoV-19/England/CAMC-B09673/2020\|EPI_ISL_673087\|20 | ···················· | ·A·················· | ····················· | ···················· | ···················· | ··A················· |
| U | 6965 | hCoV-19/England/CAMC-B097E9/2020\|EPI_ISL_673088\|20 | ···················· | ···················· | ····················· | ···················· | ···················· | ···················· |
| U | 6966 | hCoV-19/England/CAMC-B0938B/2020\|EPI_ISL_673089\|20 | ···················· | ···················· | ····················· | ···················· | ···················· | ···················· |
| U | 6967 | hCoV-19/England/CAMC-B09075/2020\|EPI_ISL_673090\|20 | ···················· | ···················· | ····················· | ···················· | ···················· | ···················· |
| U | 6968 | hCoV-19/England/CAMC-B09E20/2020\|EPI_ISL_673091\|20 | ···················· | ···················· | ····················· | ···················· | ···················· | ···················· |
| U | 6969 | hCoV-19/England/CAMC-B09312/2020\|EPI_ISL_673092\|20 | ···················· | ···················· | ····················· | ···················· | ···················· | ···················· |
| U | 6970 | hCoV-19/England/CAMC-B099A7/2020\|EPI_ISL_673093\|20 | ···················· | ···················· | ····················· | ···················· | ···················· | ···················· |
| U | 6971 | hCoV-19/England/CAMC-B09DF6/2020\|EPI_ISL_673094\|20 | ···················· | ···················· | ····················· | ···················· | ···················· | ···················· |
| U | 6972 | hCoV-19/England/CAMC-B0951C/2020\|EPI_ISL_673095\|20 | ···················· | ···················· | ····················· | ···················· | ···················· | ···················· |
| U | 6973 | hCoV-19/England/CAMC-B09145/2020\|EPI_ISL_673098\|20 | ···················· | ···················· | ····················· | ···················· | ···················· | ···················· |
| U | 6974 | hCoV-19/England/CAMC-B08E30/2020\|EPI_ISL_673100\|20 | ···················· | ···················· | ····················· | ···················· | ···················· | ···················· |
| U | 6975 | hCoV-19/England/CAMC-B08FB5/2020\|EPI_ISL_673101\|20 | ···················· | ···················· | ····················· | ···················· | ···················· | ···················· |
| U | 6976 | hCoV-19/England/CAMC-B0A00B/2020\|EPI_ISL_673103\|20 | ···················· | ·A·················· | ····················· | ···················· | ···················· | ··A················· |
| U | 6977 | hCoV-19/England/CAMC-B08E03/2020\|EPI_ISL_673104\|20 | ···················· | ···················· | ····················· | ···················· | ···················· | ···················· |
| U | 6978 | hCoV-19/England/CAMC-B094C4/2020\|EPI_ISL_673105\|20 | ···················· | ···················· | ····················· | ···················· | ···················· | ···················· |
| U | 6979 | hCoV-19/England/CAMC-B09321/2020\|EPI_ISL_673106\|20 | ···················· | ···················· | ····················· | ···················· | ···················· | ···················· |
| U | 6980 | hCoV-19/England/CAMC-B09D05/2020\|EPI_ISL_673107\|20 | ···················· | ···················· | ····················· | ···················· | ···················· | ···················· |
| U | 6981 | hCoV-19/England/CAMC-B09989/2020\|EPI_ISL_673108\|20 | ···················· | ···················· | ····················· | ···················· | ···················· | ···················· |
| U | 6982 | hCoV-19/England/CAMC-B0935E/2020\|EPI_ISL_673109\|20 | ···················· | ···················· | ····················· | ···················· | ···················· | ···················· |
| U | 6983 | hCoV-19/England/CAMC-B09EF3/2020\|EPI_ISL_673110\|20 | ···················· | ···················· | ····················· | ···················· | ···················· | ···················· |
| U | 6984 | hCoV-19/England/CAMC-B08B57/2020\|EPI_ISL_673111\|20 | ···················· | ···················· | ····················· | ···················· | ···················· | ···················· |
| U | 6985 | hCoV-19/England/CAMC-B09C62/2020\|EPI_ISL_673112\|20 | ···················· | ···················· | ····················· | ···················· | ···················· | ···················· |
| U | 6986 | hCoV-19/England/CAMC-B08E4F/2020\|EPI_ISL_673113\|20 | ···················· | ···················· | ····················· | ···················· | ···················· | ···················· |
| U | 6987 | hCoV-19/England/CAMC-B091EB/2020\|EPI_ISL_673115\|20 | ···················· | ···················· | ····················· | ···················· | ···················· | ···················· |
| U | 6988 | hCoV-19/England/CAMC-B090C0/2020\|EPI_ISL_673116\|20 | ···················· | ···················· | ····················· | ···················· | ···················· | ···················· |
| U | 6989 | hCoV-19/England/CAMC-B0988C/2020\|EPI_ISL_673117\|20 | ···················· | ···················· | ····················· | ···················· | ···················· | ···················· |
| U | 6990 | hCoV-19/England/CAMC-B0946A/2020\|EPI_ISL_673118\|20 | ···················· | ···················· | ····················· | ···················· | ···················· | ···················· |
| U | 6991 | hCoV-19/England/CAMC-B0987D/2020\|EPI_ISL_673119\|20 | ···················· | ···················· | ····················· | ···················· | ···················· | ···················· |
| U | 6992 | hCoV-19/England/CAMC-B0A0DE/2020\|EPI_ISL_673120\|20 | ···················· | ···················· | ····················· | ···················· | ···················· | ···················· |
| U | 6993 | hCoV-19/England/CAMC-B09B0B/2020\|EPI_ISL_673121\|20 | ···················· | ···················· | ····················· | ···················· | ···················· | ···················· |
| U | 6994 | hCoV-19/England/CAMC-B08F4C/2020\|EPI_ISL_673122\|20 | ···················· | ···················· | ····················· | ···················· | ···················· | ···················· |
| U | 6995 | hCoV-19/England/CAMC-B09646/2020\|EPI_ISL_673123\|20 | ···················· | ···················· | ····················· | ···················· | ···················· | ···················· |
| U | 6996 | hCoV-19/England/CAMC-B0A3E4/2020\|EPI_ISL_673124\|20 | ···················· | ···················· | ····················· | ···················· | ···················· | ···················· |
| U | 6997 | hCoV-19/England/CAMC-B091CD/2020\|EPI_ISL_673125\|20 | ···················· | ···················· | ····················· | ···················· | ···················· | ···················· |
| U | 6998 | hCoV-19/England/CAMC-B08D51/2020\|EPI_ISL_673126\|20 | ···················· | ···················· | ····················· | ···················· | ···················· | ···················· |
| U | 6999 | hCoV-19/England/CAMC-B0A4E1/2020\|EPI_ISL_673127\|20 | ···················· | ···················· | ····················· | ···················· | ···················· | ···················· |
| U | 7000 | hCoV-19/England/CAMC-B0A5DF/2020\|EPI_ISL_673131\|20 | ···················· | ···················· | ····················· | ···················· | ···················· | ···················· |
| U | 7001 | hCoV-19/England/CAMC-B08ED6/2020\|EPI_ISL_673132\|20 | ···················· | ···················· | ····················· | ···················· | ···················· | ···················· |
| U | 7002 | hCoV-19/England/CAMC-B08D9D/2020\|EPI_ISL_673133\|20 | ···················· | ···················· | ····················· | ···················· | ···················· | ··A················· |
| U | 7003 | hCoV-19/England/CAMC-B0937C/2020\|EPI_ISL_673135\|20 | ···················· | ···················· | ····················· | ···················· | ···················· | ···················· |
| U | 7004 | hCoV-19/England/ALDP-B8519C/2020\|EPI_ISL_674357\|20 | ···················· | ·R·················· | ····················· | ···················· | ···················· | ··A················· |
| U | 7005 | hCoV-19/England/MILK-B8C2DE/2020\|EPI_ISL_674358\|20 | ···················· | ···················· | ····················· | ···················· | ···················· | ···················· |
| U | 7006 | hCoV-19/England/MILK-B88700/2020\|EPI_ISL_674359\|20 | ···················· | ···················· | ····················· | ···················· | ···················· | ···················· |
| U | 7007 | hCoV-19/England/ALDP-B83DE5/2020\|EPI_ISL_674360\|20 | ···················· | ···················· | ····················· | ···················· | ···················· | ···················· |
| U | 7008 | hCoV-19/England/ALDP-B8528A/2020\|EPI_ISL_674361\|20 | ···················· | ···················· | ····················· | ···················· | ···················· | ···················· |
| U | 7009 | hCoV-19/England/MILK-B8874C/2020\|EPI_ISL_674363\|20 | ···················· | ···················· | ····················· | ···················· | ···················· | ···················· |
| U | 7010 | hCoV-19/England/MILK-B882E1/2020\|EPI_ISL_674365\|20 | ···················· | ···················· | ····················· | ···················· | ···················· | ···················· |
| U | 7011 | hCoV-19/England/ALDP-B83F49/2020\|EPI_ISL_674366\|20 | ···················· | ···················· | ····················· | ···················· | ···················· | ···················· |
| U | 7012 | hCoV-19/England/ALDP-B85141/2020\|EPI_ISL_674367\|20 | ···················· | ·A·················· | ····················· | ···················· | ···················· | ··A················· |
| U | 7013 | hCoV-19/England/ALDP-B83987/2020\|EPI_ISL_674368\|20 | ···················· | ···················· | ····················· | ···················· | ···················· | ···················· |
| U | 7014 | hCoV-19/England/ALDP-B83F0D/2020\|EPI_ISL_674369\|20 | ···················· | ···················· | ····················· | ···················· | ···················· | ···················· |
| U | 7015 | hCoV-19/England/ALDP-B85493/2020\|EPI_ISL_674370\|20 | ···················· | ···················· | ····················· | ···················· | ···················· | ···················· |
| U | 7016 | hCoV-19/England/MILK-B8C04D/2020\|EPI_ISL_674371\|20 | ···················· | ···················· | ····················· | ···················· | ···················· | ···················· |
| U | 7017 | hCoV-19/England/ALDP-B85105/2020\|EPI_ISL_674372\|20 | ···················· | ···················· | ····················· | ···················· | ···················· | ···················· |
| U | 7018 | hCoV-19/England/MILK-B88551/2020\|EPI_ISL_674373\|20 | ···················· | ···················· | ····················· | ···················· | ···················· | ···················· |
| U | 7019 | hCoV-19/England/ALDP-B8435B/2020\|EPI_ISL_674374\|20 | ···················· | ···················· | ····················· | ···················· | ···················· | ···················· |
| U | 7020 | hCoV-19/England/ALDP-B85448/2020\|EPI_ISL_674375\|20 | ···················· | ···················· | ····················· | ···················· | ···················· | ···················· |
| U | 7021 | hCoV-19/England/ALDP-B839B4/2020\|EPI_ISL_674376\|20 | ···················· | ···················· | ····················· | ···················· | ···················· | ···················· |
| U | 7022 | hCoV-19/England/MILK-B8C3EA/2020\|EPI_ISL_674378\|20 | ···················· | ···················· | ····················· | ···················· | ···················· | ···················· |
| U | 7023 | hCoV-19/England/MILK-B8C010/2020\|EPI_ISL_674379\|20 | ···················· | ···················· | ····················· | ···················· | ···················· | ···················· |
| U | 7024 | hCoV-19/England/ALDP-B843B5/2020\|EPI_ISL_674380\|20 | ···················· | ···················· | ····················· | ···················· | ···················· | ···················· |
| U | 7025 | hCoV-19/England/MILK-B88199/2020\|EPI_ISL_674381\|20 | ···················· | ···················· | ····················· | ···················· | ···················· | ···················· |
| U | 7026 | hCoV-19/England/MILK-B8C1C2/2020\|EPI_ISL_674382\|20 | ···················· | ···················· | ····················· | ···················· | ···················· | ···················· |
| U | 7027 | hCoV-19/England/ALDP-B8441C/2020\|EPI_ISL_674383\|20 | ···················· | ···················· | ····················· | ···················· | ···················· | ···················· |
| U | 7028 | hCoV-19/England/MILK-B8C55D/2020\|EPI_ISL_674384\|20 | ···················· | ···················· | ····················· | ···················· | ···················· | ···················· |
| U | 7029 | hCoV-19/England/ALDP-B849E0/2020\|EPI_ISL_674385\|20 | ···················· | ···················· | ····················· | ···················· | ···················· | ···················· |
| U | 7030 | hCoV-19/England/MILK-B884DC/2020\|EPI_ISL_674386\|20 | ···················· | ···················· | ····················· | ···················· | ···················· | ···················· |
| U | 7031 | hCoV-19/England/MILK-B883B1/2020\|EPI_ISL_674388\|20 | ···················· | ···················· | ····················· | ···················· | ···················· | ···················· |
| U | 7032 | hCoV-19/England/ALDP-B84072/2020\|EPI_ISL_674389\|20 | ···················· | ···················· | ····················· | ···················· | ···················· | ···················· |
| U | 7033 | hCoV-19/England/ALDP-B84F84/2020\|EPI_ISL_674390\|20 | ···················· | ···················· | ····················· | ···················· | ······T············· | ···················· |

| | | | | | | | | |
|---|---|---|---|---|---|---|---|---|
| U | 7034 | hCoV-19/England/ALDP-B85527/2020\|EPI_ISL_674391\|20 | ···················· | ···················· | ····················· | ···················· | ···················· | ···················· |
| U | 7035 | hCoV-19/England/ALDP-B85590/2020\|EPI_ISL_674392\|20 | ···················· | ···················· | ····················· | ···················· | ···················· | ···················· |
| U | 7036 | hCoV-19/England/ALDP-B84F2A/2020\|EPI_ISL_674396\|20 | ···················· | ···················· | ····················· | ···················· | ···················· | ···················· |
| U | 7037 | hCoV-19/England/ALDP-B84FB1/2020\|EPI_ISL_674397\|20 | ···················· | ···················· | ····················· | ···················· | ···················· | ···················· |
| U | 7038 | hCoV-19/England/MILK-B88490/2020\|EPI_ISL_674398\|20 | ···················· | ···················· | ····················· | ···················· | ···················· | ···················· |
| U | 7039 | hCoV-19/England/ALDP-B84C05/2020\|EPI_ISL_674399\|20 | ···················· | ···················· | ····················· | ···················· | ···················· | ···················· |
| U | 7040 | hCoV-19/England/ALDP-B84ACF/2020\|EPI_ISL_674400\|20 | ···················· | ···················· | ····················· | ···················· | ···················· | ···················· |
| U | 7041 | hCoV-19/England/MILK-B8C159/2020\|EPI_ISL_674401\|20 | ···················· | ···················· | ····················· | ···················· | ···················· | ···················· |
| U | 7042 | hCoV-19/England/ALDP-B85518/2020\|EPI_ISL_674402\|20 | ···················· | ···················· | ····················· | ···················· | ···················· | ···················· |
| U | 7043 | hCoV-19/England/MILK-B8C57B/2020\|EPI_ISL_674403\|20 | ···················· | ·A·················· | ····················· | ···················· | ···················· | ···A················ |
| U | 7044 | hCoV-19/England/ALDP-B83D30/2020\|EPI_ISL_674404\|20 | ···················· | ···················· | ····················· | ···················· | ···················· | ···················· |
| U | 7045 | hCoV-19/England/MILK-B8C14A/2020\|EPI_ISL_674405\|20 | ···················· | ···················· | ····················· | ···················· | ···················· | ···················· |
| U | 7046 | hCoV-19/England/ALDP-B85114/2020\|EPI_ISL_674406\|20 | ···················· | ···················· | ····················· | ···················· | ···················· | ···················· |
| U | 7047 | hCoV-19/England/ALDP-B85536/2020\|EPI_ISL_674408\|20 | ···················· | ···················· | ····················· | ···················· | ···················· | ···················· |
| U | 7048 | hCoV-19/England/ALDP-B83DD6/2020\|EPI_ISL_674409\|20 | ···················· | ···················· | ····················· | ···················· | ···················· | ···················· |
| U | 7049 | hCoV-19/England/MILK-B8876A/2020\|EPI_ISL_674410\|20 | ···················· | ···················· | ····················· | ···················· | ···················· | ···················· |
| U | 7050 | hCoV-19/England/ALDP-B85475/2020\|EPI_ISL_674411\|20 | ···················· | ···················· | ····················· | ···················· | ···················· | ···················· |
| U | 7051 | hCoV-19/England/ALDP-B84B26/2020\|EPI_ISL_674412\|20 | ···················· | ···················· | ····················· | ···················· | ···················· | ···················· |
| U | 7052 | hCoV-19/England/ALDP-B849FF/2020\|EPI_ISL_674413\|20 | ···················· | ···················· | ····················· | ···················· | ···················· | ···················· |
| U | 7053 | hCoV-19/England/MILK-B884AF/2020\|EPI_ISL_674414\|20 | ···················· | ···················· | ····················· | ···················· | ···················· | ···················· |
| U | 7054 | hCoV-19/England/MILK-B886D6/2020\|EPI_ISL_674415\|20 | ···················· | ···················· | ····················· | ···················· | ······T············· | ···················· |
| U | 7055 | hCoV-19/England/ALDP-B84C7E/2020\|EPI_ISL_674416\|20 | ···················· | ···················· | ····················· | ···················· | ···················· | ···················· |
| U | 7056 | hCoV-19/England/MILK-B8813F/2020\|EPI_ISL_674417\|20 | ···················· | ···················· | ····················· | ···················· | ···················· | ···················· |
| U | 7057 | hCoV-19/England/ALDP-B85150/2020\|EPI_ISL_674418\|20 | ···················· | ···················· | ····················· | ···················· | ···················· | ···················· |
| U | 7058 | hCoV-19/England/ALDP-B83D5E/2020\|EPI_ISL_674421\|20 | ···················· | ···················· | ····················· | ···················· | ···················· | ···················· |
| U | 7059 | hCoV-19/England/MILK-B8C49C/2020\|EPI_ISL_674422\|20 | ···················· | ···················· | ····················· | ···················· | ···················· | ···················· |
| U | 7060 | hCoV-19/England/MILK-B8832A/2020\|EPI_ISL_674423\|20 | ···················· | ···················· | ····················· | ···················· | ······T············· | ···················· |
| U | 7061 | hCoV-19/England/ALDP-B85132/2020\|EPI_ISL_674424\|20 | ···················· | ···················· | ····················· | ···················· | ···················· | ···················· |
| U | 7062 | hCoV-19/England/MILK-B8883A/2020\|EPI_ISL_674426\|20 | ···················· | ···················· | ····················· | ···················· | ···················· | ···················· |
| U | 7063 | hCoV-19/England/MILK-B8C238/2020\|EPI_ISL_674427\|20 | ···················· | ···················· | ····················· | ···················· | ···················· | ···················· |
| U | 7064 | hCoV-19/England/ALDP-B843D3/2020\|EPI_ISL_674428\|20 | ···················· | ···················· | ····················· | ···················· | ···················· | ···················· |
| U | 7065 | hCoV-19/England/ALDP-B84C9C/2020\|EPI_ISL_674429\|20 | ···················· | ···················· | ····················· | ···················· | ···················· | ···················· |
| U | 7066 | hCoV-19/England/MILK-B886E5/2020\|EPI_ISL_674430\|20 | ···················· | ···················· | ····················· | ···················· | ······T············· | ···················· |
| U | 7067 | hCoV-19/England/ALDP-B85545/2020\|EPI_ISL_674432\|20 | ···················· | ···················· | ····················· | ···················· | ···················· | ···················· |
| U | 7068 | hCoV-19/England/MILK-B8C5A8/2020\|EPI_ISL_674433\|20 | ···················· | ···················· | ····················· | ···················· | ···················· | ···················· |
| U | 7069 | hCoV-19/England/ALDP-B84054/2020\|EPI_ISL_674434\|20 | ···················· | ···················· | ····················· | ···················· | ···················· | ···················· |
| U | 7070 | hCoV-19/England/ALDP-B8436A/2020\|EPI_ISL_674435\|20 | ···················· | ···················· | ····················· | ···················· | ···················· | ···················· |
| U | 7071 | hCoV-19/England/ALDP-B84027/2020\|EPI_ISL_674436\|20 | ···················· | ···················· | ····················· | ···················· | ···················· | ···················· |
| U | 7072 | hCoV-19/England/MILK-B8C0B6/2020\|EPI_ISL_674437\|20 | ···················· | ···················· | ····················· | ···················· | ···················· | ···················· |
| U | 7073 | hCoV-19/England/ALDP-B839C3/2020\|EPI_ISL_674438\|20 | ···················· | ···················· | ····················· | ···················· | ···················· | ···················· |
| U | 7074 | hCoV-19/England/ALDP-B85509/2020\|EPI_ISL_674439\|20 | ···················· | ·········T·········· | ····················· | ···················· | ···················· | ···················· |
| U | 7075 | hCoV-19/England/MILK-B887F1/2020\|EPI_ISL_674440\|20 | ···················· | ···················· | ····················· | ···················· | ···················· | ···················· |
| U | 7076 | hCoV-19/England/MILK-B8871F/2020\|EPI_ISL_674443\|20 | ···················· | ···················· | ····················· | ···················· | ···················· | ···················· |
| U | 7077 | hCoV-19/England/MILK-B88418/2020\|EPI_ISL_674444\|20 | ···················· | ···················· | ····················· | ···················· | ···················· | ···················· |
| U | 7078 | hCoV-19/England/ALDP-B84036/2020\|EPI_ISL_674445\|20 | ···················· | ···················· | ····················· | ···················· | ···················· | ···················· |
| U | 7079 | hCoV-19/England/ALDP-B84C6F/2020\|EPI_ISL_674446\|20 | ···················· | ···················· | ····················· | ···················· | ···················· | ···················· |
| U | 7080 | hCoV-19/England/MILK-B8C256/2020\|EPI_ISL_674448\|20 | ···················· | ·A·················· | ····················· | ···················· | ···················· | ···A················ |
| U | 7081 | hCoV-19/England/MILK-B88348/2020\|EPI_ISL_674450\|20 | ···················· | ···················· | ····················· | ···················· | ···················· | ···················· |
| U | 7082 | hCoV-19/England/ALDP-B84045/2020\|EPI_ISL_674451\|20 | ···················· | ···················· | ····················· | ···················· | ···················· | ···················· |
| U | 7083 | hCoV-19/England/MILK-B8C502/2020\|EPI_ISL_674453\|20 | ···················· | ···················· | ····················· | ···················· | ···················· | ···················· |
| U | 7084 | hCoV-19/England/ALDP-B8433D/2020\|EPI_ISL_674454\|20 | ···················· | ·A·················· | ····················· | ···················· | ···················· | ···A················ |
| U | 7085 | hCoV-19/England/ALDP-B843C4/2020\|EPI_ISL_674455\|20 | ···················· | ···················· | ····················· | ···················· | ···················· | ···················· |
| U | 7086 | hCoV-19/England/ALDP-B84A56/2020\|EPI_ISL_674456\|20 | ···················· | ···················· | ····················· | ···················· | ···················· | ···················· |
| U | 7087 | hCoV-19/England/MILK-B8C1A4/2020\|EPI_ISL_674457\|20 | ···················· | ···················· | ····················· | ···················· | ···················· | ···················· |
| U | 7088 | hCoV-19/England/MILK-B88409/2020\|EPI_ISL_674458\|20 | ···················· | ···················· | ····················· | ···················· | ···················· | ···················· |
| U | 7089 | hCoV-19/England/MILK-B8C353/2020\|EPI_ISL_674460\|20 | ···················· | ···················· | ····················· | ···················· | ···················· | ···················· |
| U | 7090 | hCoV-19/England/ALDP-B839F0/2020\|EPI_ISL_674462\|20 | ···················· | ···················· | ····················· | ···················· | ···················· | ···················· |
| U | 7091 | hCoV-19/England/MILK-B8831B/2020\|EPI_ISL_674464\|20 | ···················· | ···················· | ····················· | ···················· | ···················· | ···················· |
| U | 7092 | hCoV-19/England/ALDP-B83D6D/2020\|EPI_ISL_674465\|20 | ···················· | ···················· | ····················· | ···················· | ···················· | ···················· |
| U | 7093 | hCoV-19/England/ALDP-B85387/2020\|EPI_ISL_675688\|20 | ···················· | ···················· | ····················· | ···················· | ···················· | ···················· |
| U | 7094 | hCoV-19/England/ALDP-B85ACE/2020\|EPI_ISL_675689\|20 | ···················· | ···················· | ····················· | ···················· | ···················· | ···················· |
| U | 7095 | hCoV-19/England/ALDP-B8584C/2020\|EPI_ISL_675691\|20 | ···················· | ···················· | ····················· | ···················· | ···················· | ···················· |
| U | 7096 | hCoV-19/England/ALDP-B83565/2020\|EPI_ISL_675692\|20 | ···················· | ·A·················· | ····················· | ···················· | ···················· | ···A················ |
| U | 7097 | hCoV-19/England/MILK-B89CA6/2020\|EPI_ISL_675693\|20 | ···················· | ·A·················· | ····················· | ···················· | ···················· | ···A················ |
| U | 7098 | hCoV-19/England/MILK-B89C1F/2020\|EPI_ISL_675694\|20 | ···················· | ···················· | ····················· | ···················· | ···················· | ···················· |
| U | 7099 | hCoV-19/England/ALDP-B85B34/2020\|EPI_ISL_675696\|20 | ···················· | ···················· | ····················· | ···················· | ···················· | ···················· |
| U | 7100 | hCoV-19/England/ALDP-B85958/2020\|EPI_ISL_675697\|20 | ···················· | ···················· | ····················· | ···················· | ···················· | ···················· |
| U | 7101 | hCoV-19/England/MILK-B89C2E/2020\|EPI_ISL_675698\|20 | ···················· | ···················· | ····················· | ···················· | ···················· | ···················· |
| U | 7102 | hCoV-19/England/ALDP-B85378/2020\|EPI_ISL_675699\|20 | ···················· | ···················· | ····················· | ···················· | ···················· | ···················· |
| U | 7103 | hCoV-19/England/MILK-B89778/2020\|EPI_ISL_675722\|20 | ···················· | ···················· | ····················· | ···················· | ······T············· | ···················· |
| U | 7104 | hCoV-19/England/MILK-B8C7DF/2020\|EPI_ISL_675723\|20 | ···················· | ···················· | ····················· | ···················· | ···················· | ···················· |
| U | 7105 | hCoV-19/England/MILK-B88BAA/2020\|EPI_ISL_675724\|20 | ···················· | ·A·················· | ····················· | ···················· | ···················· | ···A················ |
| U | 7106 | hCoV-19/England/MILK-B8C687/2020\|EPI_ISL_675725\|20 | ···················· | ···················· | ····················· | ···················· | ···················· | ···················· |
| U | 7107 | hCoV-19/England/MILK-B8CA03/2020\|EPI_ISL_675726\|20 | ···················· | ···················· | ····················· | ···················· | ···················· | ···················· |
| U | 7108 | hCoV-19/England/MILK-B8C70C/2020\|EPI_ISL_675729\|20 | ···················· | ···················· | ····················· | ···················· | ···················· | ···················· |
| U | 7109 | hCoV-19/England/MILK-B895F6/2020\|EPI_ISL_675731\|20 | ···················· | ···················· | ····················· | ···················· | ···················· | ···················· |
| U | 7110 | hCoV-19/England/MILK-B895D8/2020\|EPI_ISL_675732\|20 | ···················· | ···················· | ····················· | ···················· | ···················· | ···················· |
| U | 7111 | hCoV-19/England/MILK-B8C4F6/2020\|EPI_ISL_675734\|20 | ···················· | ···················· | ····················· | ···················· | ···················· | ···················· |
| U | 7112 | hCoV-19/England/ALDP-B84D3F/2020\|EPI_ISL_675735\|20 | ···················· | ···················· | ····················· | ···················· | ···················· | ···················· |
| U | 7113 | hCoV-19/England/MILK-B897C3/2020\|EPI_ISL_675737\|20 | ···················· | ···················· | ····················· | ···················· | ···················· | ···················· |

| | | | | | | | | |
|---|---|---|---|---|---|---|---|---|
| U | 7114 hCoV-19/England/ALDP-B85985/2020\|EPI_ISL_675740\|20 | ···················· | ···················· | ···················· | ···················· | ···················· | ···················· | |
| U | 7115 hCoV-19/England/MILK-B88BD7/2020\|EPI_ISL_675741\|20 | ···················· | ···················· | ···················· | ···················· | ···················· | ···················· | |
| U | 7116 hCoV-19/England/MILK-B88D2C/2020\|EPI_ISL_675743\|20 | ···················· | ···················· | ···················· | ···················· | ···················· | ···················· | |
| U | 7117 hCoV-19/England/ALDP-B85A19/2020\|EPI_ISL_675746\|20 | ···················· | ···················· | ···················· | ···················· | ···················· | ···················· | |
| U | 7118 hCoV-19/England/MILK-B88A70/2020\|EPI_ISL_675747\|20 | ···················· | ···················· | ···················· | ···················· | ···················· | ···················· | |
| U | 7119 hCoV-19/England/MILK-B88991/2020\|EPI_ISL_675749\|20 | ···················· | ···················· | ···················· | ···················· | ···················· | ···················· | |
| U | 7120 hCoV-19/England/ALDP-B8483E/2020\|EPI_ISL_675750\|20 | ···················· | ···················· | ···················· | ···················· | ···················· | ···················· | |
| U | 7121 hCoV-19/England/ALDP-B85071/2020\|EPI_ISL_675751\|20 | ···················· | ···················· | ···················· | ···················· | ···················· | ···················· | |
| U | 7122 hCoV-19/England/MILK-B8C881/2020\|EPI_ISL_675752\|20 | ···················· | ···················· | ···················· | ···················· | ···················· | ···················· | |
| U | 7123 hCoV-19/England/MILK-B897B4/2020\|EPI_ISL_675753\|20 | ···················· | ···················· | ···················· | ···················· | ···················· | ···················· | |
| U | 7124 hCoV-19/England/MILK-B8C7C0/2020\|EPI_ISL_675754\|20 | ···················· | ···················· | ···················· | ···················· | ···················· | ···················· | |
| U | 7125 hCoV-19/England/MILK-B8882B/2020\|EPI_ISL_675755\|20 | ···················· | ···················· | ···················· | ···················· | ···················· | ···················· | |
| U | 7126 hCoV-19/England/ALDP-B859C1/2020\|EPI_ISL_675756\|20 | ···················· | ···················· | ···················· | ···················· | ···················· | ···················· | |
| U | 7127 hCoV-19/England/ALDP-B85053/2020\|EPI_ISL_675757\|20 | ···················· | ···················· | ···················· | ···················· | ···················· | ···················· | |
| U | 7128 hCoV-19/England/ALDP-B85A0A/2020\|EPI_ISL_675759\|20 | ···················· | ·A·················· | ···················· | ···················· | ···················· | ··A················· | |
| U | 7129 hCoV-19/England/ALDP-B859D0/2020\|EPI_ISL_675760\|20 | ···················· | ···················· | ···················· | ···················· | ···················· | ···················· | |
| U | 7130 hCoV-19/England/MILK-B8C64B/2020\|EPI_ISL_675762\|20 | ···················· | ···················· | ···················· | ···················· | ···················· | ···················· | |
| U | 7131 hCoV-19/England/MILK-B8C942/2020\|EPI_ISL_675763\|20 | ···················· | ···················· | ···················· | ···················· | ···················· | ···················· | |
| U | 7132 hCoV-19/England/MILK-B8C6B4/2020\|EPI_ISL_675765\|20 | ···················· | ···················· | ···················· | ···················· | ···················· | ···················· | |
| U | 7133 hCoV-19/England/ALDP-B848C5/2020\|EPI_ISL_675766\|20 | ···················· | ···················· | ···················· | ···················· | ···················· | ···················· | |
| U | 7134 hCoV-19/England/ALDP-B84EA5/2020\|EPI_ISL_675771\|20 | ···················· | ···················· | ···················· | ···················· | ···················· | ···················· | |
| U | 7135 hCoV-19/England/MILK-B8C54E/2020\|EPI_ISL_675772\|20 | ···················· | ···················· | ···················· | ···················· | ···················· | ···················· | |
| U | 7136 hCoV-19/England/ALDP-B850BD/2020\|EPI_ISL_675774\|20 | ···················· | ···················· | ···················· | ···················· | ···················· | ···················· | |
| U | 7137 hCoV-19/England/ALDP-B85026/2020\|EPI_ISL_675775\|20 | ···················· | ···················· | ···················· | ···················· | ···················· | ···················· | |
| U | 7138 hCoV-19/England/ALDP-B8475F/2020\|EPI_ISL_675776\|20 | ···················· | ···················· | ···················· | ···················· | ···················· | ···················· | |
| U | 7139 hCoV-19/England/ALDP-B84DE4/2020\|EPI_ISL_675777\|20 | ···················· | ···················· | ···················· | ···················· | ···················· | ···················· | |
| U | 7140 hCoV-19/England/ALDP-B849C2/2020\|EPI_ISL_675779\|20 | ···················· | ···················· | ···················· | ···················· | ···················· | ···················· | |
| U | 7141 hCoV-19/England/MILK-B8CB6A/2020\|EPI_ISL_675781\|20 | ···················· | ···················· | ···················· | ···················· | ···················· | ···················· | |
| U | 7142 hCoV-19/England/MILK-B8CAA9/2020\|EPI_ISL_675783\|20 | ···················· | ···················· | ···················· | ···················· | ···················· | ···················· | |
| U | 7143 hCoV-19/England/MILK-B8C9AC/2020\|EPI_ISL_675784\|20 | ···················· | ···················· | ···················· | ···················· | ···················· | ···················· | |
| U | 7144 hCoV-19/England/ALDP-B849B3/2020\|EPI_ISL_675785\|20 | ···················· | ···················· | ···················· | ···················· | ···················· | ···················· | |
| U | 7145 hCoV-19/England/MILK-B8CA6D/2020\|EPI_ISL_675786\|20 | ···················· | ···················· | ···················· | ···················· | ···················· | ···················· | |
| U | 7146 hCoV-19/England/MILK-B8C4BA/2020\|EPI_ISL_675787\|20 | ···················· | ···················· | ···················· | ···················· | ···················· | ···················· | |
| U | 7147 hCoV-19/England/ALDP-B85008/2020\|EPI_ISL_675788\|20 | ···················· | ···················· | ···················· | ···················· | ···················· | ···················· | |
| U | 7148 hCoV-19/England/MILK-B88D3B/2020\|EPI_ISL_675795\|20 | ···················· | ···················· | ···················· | ···················· | ···················· | ···················· | |
| U | 7149 hCoV-19/England/CAMC-BBC86A/2020\|EPI_ISL_679249\|20 | ···················· | ···················· | ···················· | ···················· | ···················· | ···················· | |
| U | 7151 hCoV-19/Northern Ireland/NIRE-109798/2020\|EPI_ISL_ | ···················· | ···················· | ···················· | ···················· | ···················· | ···················· | |
| U | 7152 hCoV-19/Northern Ireland/NIRE-232E63/2020\|EPI_ISL_ | ···················· | ···················· | ···················· | ···················· | ···················· | ···················· | |
| U | 7153 hCoV-19/England/MILK-BAA674/2020\|EPI_ISL_681551\|20 | ···················· | ·A·················· | ···················· | ···················· | ···················· | ···················· | |
| U | 7154 hCoV-19/England/MILK-BA9E22/2020\|EPI_ISL_681555\|20 | ···················· | ···················· | ···················· | ···················· | ···················· | ···················· | |
| U | 7155 hCoV-19/England/MILK-BAA39B/2020\|EPI_ISL_681556\|20 | ···················· | ···················· | ···················· | ···················· | ···················· | ···················· | |
| U | 7156 hCoV-19/England/MILK-BA9AB5/2020\|EPI_ISL_681557\|20 | ···················· | ···················· | ···················· | ···················· | ···················· | ···················· | |
| U | 7157 hCoV-19/England/MILK-BA9BD0/2020\|EPI_ISL_681559\|20 | ···················· | ···················· | ···················· | ···················· | ···················· | ···················· | |
| U | 7158 hCoV-19/England/MILK-BAA37D/2020\|EPI_ISL_681560\|20 | ···················· | ···················· | ···················· | ···················· | ···················· | ···················· | |
| U | 7159 hCoV-19/Denmark/DCGC-12619/2020\|EPI_ISL_682800\|202 | ···················· | ···················· | ···················· | ···················· | ···················· | ···················· | |
| U | 7160 hCoV-19/Denmark/DCGC-12620/2020\|EPI_ISL_682801\|202 | ···················· | ···················· | ···················· | ···················· | ···················· | ···················· | |
| U | 7161 hCoV-19/Denmark/DCGC-12621/2020\|EPI_ISL_682802\|202 | ···················· | ···················· | ···················· | ···················· | ···················· | ···················· | |
| U | 7162 hCoV-19/Denmark/DCGC-12622/2020\|EPI_ISL_682803\|202 | ···················· | ···················· | ···················· | ···················· | ···················· | ···················· | |
| U | 7163 hCoV-19/Denmark/DCGC-12623/2020\|EPI_ISL_682804\|202 | ···················· | ···················· | ···················· | ···················· | ···················· | ···················· | |
| U | 7164 hCoV-19/Denmark/DCGC-12624/2020\|EPI_ISL_682805\|202 | ···················· | ···················· | ···················· | ···················· | ···················· | ···················· | |
| U | 7165 hCoV-19/Denmark/DCGC-12625/2020\|EPI_ISL_682806\|202 | ···················· | ···················· | ···················· | ···················· | ···················· | ···················· | |
| U | 7166 hCoV-19/Denmark/DCGC-12628/2020\|EPI_ISL_682809\|202 | ···················· | ···················· | ···················· | ···················· | ······T············· | ···················· | |
| U | 7167 hCoV-19/Denmark/DCGC-12705/2020\|EPI_ISL_682886\|202 | ···················· | ···················· | ···················· | ···················· | ···················· | ···················· | |
| U | 7168 hCoV-19/Denmark/DCGC-12711/2020\|EPI_ISL_682892\|202 | ···················· | ···················· | ···················· | ···················· | ···················· | ···················· | |
| U | 7169 hCoV-19/USA/NY-Wadsworth-76683-01/2020\|EPI_ISL_676 | ···················· | ···················· | ···················· | ···················· | ···················· | ···················· | |
| U | 7170 hCoV-19/USA/NY-Wadsworth-76695-01/2020\|EPI_ISL_676 | ···················· | ···················· | ···················· | ···················· | ···················· | ···················· | |
| U | 7171 hCoV-19/USA/NY-Wadsworth-76829-01/2020\|EPI_ISL_676 | ···················· | ···················· | ···················· | ···················· | ···················· | ···················· | |
| U | 7172 hCoV-19/USA/NY-Wadsworth-77659-01/2020\|EPI_ISL_676 | ···················· | ···················· | ···················· | ···················· | ···················· | ···················· | |
| U | 7173 hCoV-19/USA/NY-Wadsworth-77837-01/2020\|EPI_ISL_676 | ···················· | ···················· | ···················· | ···················· | ···················· | ···················· | |
| U | 7174 hCoV-19/USA/NY-Wadsworth-78727-01/2020\|EPI_ISL_676 | ···················· | ···················· | ···················· | ···················· | ···················· | ···················· | |
| U | 7175 hCoV-19/USA/NY-Wadsworth-78728-01/2020\|EPI_ISL_676 | ···················· | ···················· | ···················· | ···················· | ···················· | ···················· | |
| U | 7176 hCoV-19/USA/NY-Wadsworth-79524-01/2020\|EPI_ISL_676 | ···················· | ···················· | ···················· | ···················· | ···················· | ···················· | |
| U | 7177 hCoV-19/USA/NY-Wadsworth-79575-01/2020\|EPI_ISL_676 | ···················· | ···················· | ···················· | ···················· | ···················· | ···················· | |
| U | 7178 hCoV-19/USA/NY-Wadsworth-79894-01/2020\|EPI_ISL_676 | ···················· | ···················· | ···················· | ···················· | ···················· | ···················· | |
| U | 7179 hCoV-19/USA/NY-Wadsworth-79903-01/2020\|EPI_ISL_676 | ···················· | ···················· | ···················· | ···················· | ···················· | ···················· | |
| U | 7180 hCoV-19/USA/NY-Wadsworth-80578-01/2020\|EPI_ISL_676 | ···················· | ···················· | ···················· | ···················· | ···················· | ···················· | |
| U | 7181 hCoV-19/USA/NY-Wadsworth-80638-01/2020\|EPI_ISL_676 | ···················· | ···················· | ···················· | ···················· | ···················· | ···················· | |
| U | 7182 hCoV-19/USA/NY-Wadsworth-81196-01/2020\|EPI_ISL_676 | ···················· | ···················· | ···················· | ···················· | ···················· | ···················· | |
| U | 7183 hCoV-19/USA/NY-Wadsworth-81540-01/2020\|EPI_ISL_676 | ···················· | ···················· | ···················· | ···················· | ·················C·· | ···················· | 1 |
| U | 7184 hCoV-19/USA/NY-Wadsworth-83699-01/2020\|EPI_ISL_676 | ···················· | ···················· | ···················· | ···················· | ···················· | ···················· | |
| U | 7185 hCoV-19/USA/NY-Wadsworth-88095-01/2020\|EPI_ISL_676 | ···················· | ···················· | ···················· | ···················· | ···················· | ···················· | |
| U | 7186 hCoV-19/USA/NY-Wadsworth-88316-01/2020\|EPI_ISL_676 | ···················· | ···················· | ···················· | ···················· | ···················· | ···················· | |
| U | 7187 hCoV-19/USA/NY-Wadsworth-88325-01/2020\|EPI_ISL_676 | ···················· | ···················· | ···················· | ···················· | ···················· | ···················· | |
| U | 7188 hCoV-19/USA/NY-Wadsworth-88367-01/2020\|EPI_ISL_676 | ···················· | ···················· | ···················· | ···················· | ···················· | ···················· | |
| U | 7189 hCoV-19/USA/NY-Wadsworth-95277-01/2020\|EPI_ISL_676 | ···················· | ···················· | ···················· | ···················· | ···················· | ···················· | |
| U | 7190 hCoV-19/USA/NY-Wadsworth-95907-01/2020\|EPI_ISL_676 | ···················· | ···················· | ···················· | ···················· | ···················· | ···················· | |
| U | 7191 hCoV-19/USA/NY-Wadsworth-96361-01/2020\|EPI_ISL_676 | ···················· | ···················· | ···················· | ···················· | ···················· | ···················· | |
| U | 7192 hCoV-19/USA/NY-Wadsworth-96601-01/2020\|EPI_ISL_676 | ···················· | ···················· | ···················· | ···················· | ···················· | ···················· | |
| U | 7193 hCoV-19/USA/NY-Wadsworth-97721-01/2020\|EPI_ISL_676 | ···················· | ···················· | ···················· | ···················· | ···················· | ···················· | |
| U | 7194 hCoV-19/USA/NY-Wadsworth-97726-01/2020\|EPI_ISL_676 | ···················· | ···················· | ···················· | ···················· | ···················· | ···················· | |

| | | | | | | | | |
|---|---|---|---|---|---|---|---|---|
| U | 7195 | hCoV-19/USA/NY-Wadsworth-97804-01/2020\|EPI_ISL_676 | ···················· | ···················· | ···················· | ···················· | ···················· | ···················· |
| U | 7196 | hCoV-19/USA/NY-Wadsworth-97827-01/2020\|EPI_ISL_676 | ···················· | ···················· | ···················· | ···················· | ···················· | ···················· |
| U | 7197 | hCoV-19/USA/NY-Wadsworth-97869-01/2020\|EPI_ISL_676 | ···················· | ···················· | ···················· | ···················· | ···················· | ···················· |
| U | 7198 | hCoV-19/USA/NY-Wadsworth-97873-01/2020\|EPI_ISL_676 | ···················· | ···················· | ···················· | ···················· | ···················· | ···················· |
| U | 7199 | hCoV-19/USA/NY-Wadsworth-99255-01/2020\|EPI_ISL_676 | ···················· | ···················· | ···················· | ···················· | ···················· | ···················· |
| U | 7200 | hCoV-19/England/BIRM-6C03F/2020\|EPI_ISL_679130\|202 | ···················· | ········A··········· | ···················· | ···················· | ···················· | ···················· |
| U | 7201 | hCoV-19/England/BIRM-6C04E/2020\|EPI_ISL_679131\|202 | ···················· | ···················· | ···················· | ···················· | ···················· | ···················· |
| U | 7202 | hCoV-19/England/BIRM-6C06C/2020\|EPI_ISL_679132\|202 | ···················· | ···················· | ···················· | ···················· | ···················· | ···················· |
| U | 7203 | hCoV-19/England/CAMB-1B93FD/2020\|EPI_ISL_679134\|20 | ···················· | ···················· | ···················· | ···················· | ···················· | ···················· |
| U | 7204 | hCoV-19/England/CAMB-1B9515/2020\|EPI_ISL_679137\|20 | ···················· | ···················· | ···················· | ···················· | ···················· | ···················· |
| U | 7205 | hCoV-19/England/CAMB-1B9533/2020\|EPI_ISL_679138\|20 | ···················· | ···················· | ···················· | ···················· | ···················· | ···················· |
| U | 7206 | hCoV-19/England/CAMB-1B983A/2020\|EPI_ISL_679143\|20 | ···················· | ···················· | ···················· | ···················· | ···················· | ···················· |
| U | 7207 | hCoV-19/England/CAMB-1B9A52/2020\|EPI_ISL_679146\|20 | ···················· | ···················· | ···················· | ···················· | ···················· | ···················· |
| U | 7208 | hCoV-19/England/CAMB-1B9A61/2020\|EPI_ISL_679147\|20 | ···················· | ···················· | ···················· | ···················· | ·····T·············· | ···················· |
| U | 7209 | hCoV-19/England/CAMB-1B9ACB/2020\|EPI_ISL_679149\|20 | ···················· | ·A·················· | ···················· | ···················· | ···················· | ··A················· |
| U | 7210 | hCoV-19/England/CAMB-1B9B22/2020\|EPI_ISL_679152\|20 | ···················· | ···················· | ···················· | ···················· | ···················· | ···················· |
| U | 7211 | hCoV-19/England/CAMB-1B9B40/2020\|EPI_ISL_679153\|20 | ···················· | ···················· | ···················· | ···················· | ···················· | ···················· |
| U | 7212 | hCoV-19/England/CAMB-1BA31A/2020\|EPI_ISL_679155\|20 | ···················· | ···················· | ···················· | ···················· | ···················· | ···················· |
| U | 7213 | hCoV-19/England/CAMB-1BA505/2020\|EPI_ISL_679156\|20 | ···················· | ···················· | ···················· | ···················· | ···················· | ···················· |
| U | 7214 | hCoV-19/England/CAMB-1BA5D8/2020\|EPI_ISL_679157\|20 | ···················· | ···················· | ···················· | ···················· | ···················· | ···················· |
| U | 7215 | hCoV-19/England/CAMB-1BA611/2020\|EPI_ISL_679158\|20 | ···················· | ···················· | ···················· | ···················· | ···················· | ···················· |
| U | 7216 | hCoV-19/England/CAMB-1BB908/2020\|EPI_ISL_679161\|20 | ···················· | ···················· | ···················· | ···················· | ···················· | ···················· |
| U | 7217 | hCoV-19/England/CAMB-1BB917/2020\|EPI_ISL_679162\|20 | ···················· | ···················· | ···················· | ···················· | ···················· | ···················· |
| U | 7218 | hCoV-19/England/CAMB-1BB935/2020\|EPI_ISL_679164\|20 | ···················· | ···················· | ···················· | ···················· | ···················· | ···················· |
| U | 7219 | hCoV-19/England/CAMB-1BB962/2020\|EPI_ISL_679167\|20 | ···················· | ···················· | ···················· | ···················· | ···················· | ···················· |
| U | 7220 | hCoV-19/England/CAMB-1BB971/2020\|EPI_ISL_679168\|20 | ···················· | ···················· | ···················· | ···················· | ···················· | ···················· |
| U | 7221 | hCoV-19/England/CAMB-1BB980/2020\|EPI_ISL_679169\|20 | ···················· | ···················· | ···················· | ···················· | ···················· | ···················· |
| U | 7222 | hCoV-19/England/CAMB-1BB99F/2020\|EPI_ISL_679170\|20 | ···················· | ···················· | ···················· | ···················· | ·····T·············· | ···················· |
| U | 7223 | hCoV-19/England/CAMB-1BB9DB/2020\|EPI_ISL_679174\|20 | ···················· | ···················· | ···················· | ···················· | ···················· | ···················· |
| U | 7224 | hCoV-19/England/CAMB-1BBA14/2020\|EPI_ISL_679178\|20 | ···················· | ···················· | ···················· | ···················· | ·····T·············· | ···················· |
| U | 7225 | hCoV-19/England/CAMB-1BBA23/2020\|EPI_ISL_679179\|20 | ···················· | ···················· | ···················· | ···················· | ·····T·············· | ···················· |
| U | 7226 | hCoV-19/England/CAMB-1BBA32/2020\|EPI_ISL_679180\|20 | ···················· | ···················· | ···················· | ···················· | ···················· | ···················· |
| U | 7227 | hCoV-19/England/CAMB-1BBA50/2020\|EPI_ISL_679182\|20 | ···················· | ···················· | ···················· | ···················· | ···················· | ···················· |
| U | 7228 | hCoV-19/England/CAMB-1BBA6F/2020\|EPI_ISL_679183\|20 | ···················· | ···················· | ···················· | ···················· | ·····T·············· | ···················· |
| U | 7229 | hCoV-19/England/CAMB-1BBA9C/2020\|EPI_ISL_679186\|20 | ···················· | ···················· | ···················· | ···················· | ···················· | ···················· |
| U | 7230 | hCoV-19/England/CAMB-1BBAAB/2020\|EPI_ISL_679187\|20 | ···················· | ···················· | ···················· | ···················· | ···················· | ···················· |
| U | 7231 | hCoV-19/England/CAMB-1BBABA/2020\|EPI_ISL_679188\|20 | ···················· | ···················· | ···················· | ···················· | ·····T·············· | ···················· |
| U | 7232 | hCoV-19/England/CAMB-1BBAC9/2020\|EPI_ISL_679189\|20 | ···················· | ···················· | ···················· | ···················· | ···················· | ···················· |
| U | 7233 | hCoV-19/England/CAMB-1BBAE7/2020\|EPI_ISL_679191\|20 | ···················· | ···················· | ···················· | ···················· | ···················· | ···················· |
| U | 7234 | hCoV-19/England/CAMB-1BBB02/2020\|EPI_ISL_679193\|20 | ···················· | ···················· | ···················· | ···················· | ···················· | ···················· |
| U | 7235 | hCoV-19/England/CAMB-1BBB3F/2020\|EPI_ISL_679196\|20 | ···················· | ···················· | ···················· | ···················· | ···················· | ···················· |
| U | 7236 | hCoV-19/England/CAMB-1BBB5D/2020\|EPI_ISL_679198\|20 | ···················· | ···················· | ···················· | ···················· | ···················· | ···················· |
| U | 7237 | hCoV-19/England/CAMB-1BBC69/2020\|EPI_ISL_679214\|20 | ···················· | ···················· | ···················· | ···················· | ···················· | ···················· |
| U | 7238 | hCoV-19/England/CAMB-1BBD2A/2020\|EPI_ISL_679224\|20 | ···················· | ···················· | ···················· | ···················· | ···················· | ···················· |
| U | 7239 | hCoV-19/England/SHEF-C17BB/2020\|EPI_ISL_680118\|202 | ···················· | ···················· | ···················· | ···················· | ···················· | ···················· |
| U | 7240 | hCoV-19/England/SHEF-C2037/2020\|EPI_ISL_680120\|202 | ···················· | ···················· | ···················· | ···················· | ···················· | ···················· |
| U | 7241 | hCoV-19/England/SHEF-C236B/2020\|EPI_ISL_680121\|202 | ···················· | ···················· | ···················· | ···················· | ···················· | ···················· |
| U | 7242 | hCoV-19/England/SHEF-C2DD6/2020\|EPI_ISL_680126\|202 | ···················· | ···················· | ···················· | ···················· | ···················· | ···················· |
| U | 7243 | hCoV-19/England/SHEF-C2EE2/2020\|EPI_ISL_680127\|202 | ···················· | ···················· | ···················· | ···················· | ···················· | ···················· |
| U | 7244 | hCoV-19/England/SHEF-C38E3/2020\|EPI_ISL_680129\|202 | ···················· | ···················· | ···················· | ···················· | ···················· | ···················· |
| U | 7245 | hCoV-19/England/SHEF-C431E/2020\|EPI_ISL_680134\|202 | ···················· | ···················· | ···················· | ···················· | ···················· | ···················· |
| U | 7246 | hCoV-19/England/SHEF-C46F7/2020\|EPI_ISL_680135\|202 | ···················· | ···················· | ···················· | ···················· | ···················· | ···················· |
| U | 7247 | hCoV-19/England/SHEF-C5B8E/2020\|EPI_ISL_680142\|202 | ···················· | ···················· | ···················· | ···················· | ···················· | ···················· |
| U | 7248 | hCoV-19/England/SHEF-C669B/2020\|EPI_ISL_680144\|202 | ···················· | ···················· | ···················· | ···················· | ···················· | ···················· |
| U | 7249 | hCoV-19/England/SHEF-C7D95/2020\|EPI_ISL_680149\|202 | ···················· | ···················· | ···················· | ···················· | ···················· | ···················· |
| U | 7250 | hCoV-19/England/SHEF-C8E73/2020\|EPI_ISL_680157\|202 | ···················· | ···················· | ···················· | ···················· | ···················· | ···················· |
| U | 7251 | hCoV-19/England/SHEF-C9285/2020\|EPI_ISL_680158\|202 | ···················· | ···················· | ···················· | ···················· | ···················· | ···················· |
| U | 7252 | hCoV-19/England/SHEF-C959B/2020\|EPI_ISL_680160\|202 | ···················· | ···················· | ···················· | ···················· | ···················· | ···················· |
| U | 7253 | hCoV-19/England/SHEF-C9E90/2020\|EPI_ISL_680163\|202 | ···················· | ···················· | ···················· | ···················· | ···················· | ···················· |
| U | 7254 | hCoV-19/England/SHEF-CA318/2020\|EPI_ISL_680165\|202 | ···················· | ···················· | ···················· | ···················· | ···················· | ···················· |
| U | 7255 | hCoV-19/England/SHEF-CA5D6/2020\|EPI_ISL_680167\|202 | ···················· | ···················· | ···················· | ···················· | ···················· | ···················· |
| U | 7256 | hCoV-19/England/SHEF-CAAE6/2020\|EPI_ISL_680169\|202 | ···················· | ···················· | ···················· | ···················· | ···················· | ···················· |
| U | 7257 | hCoV-19/England/SHEF-CB265/2020\|EPI_ISL_680170\|202 | ···················· | ···················· | ···················· | ···················· | ···················· | ···················· |
| U | 7258 | hCoV-19/England/SHEF-CBB4C/2020\|EPI_ISL_680175\|202 | ···················· | ···················· | ···················· | ···················· | ···················· | ···················· |
| U | 7259 | hCoV-19/England/SHEF-CBC49/2020\|EPI_ISL_680176\|202 | ···················· | ···················· | ···················· | ···················· | ···················· | ···················· |
| U | 7260 | hCoV-19/England/SHEF-CC871/2020\|EPI_ISL_680179\|202 | ···················· | ···················· | ···················· | ···················· | ···················· | ···················· |
| U | 7261 | hCoV-19/England/SHEF-CCA8A/2020\|EPI_ISL_680181\|202 | ···················· | ···················· | ···················· | ···················· | ···················· | ···················· |
| U | 7262 | hCoV-19/England/SHEF-CF6CF/2020\|EPI_ISL_680186\|202 | ···················· | ···················· | ···················· | ···················· | ···················· | ···················· |
| U | 7263 | hCoV-19/England/SHEF-CFEC7/2020\|EPI_ISL_680188\|202 | ···················· | ···················· | ···················· | ···················· | ···················· | ···················· |
| U | 7264 | hCoV-19/England/SHEF-D0476/2020\|EPI_ISL_680189\|202 | ···················· | ···················· | ···················· | ···················· | ···················· | ···················· |
| U | 7265 | hCoV-19/England/SHEF-D14C0/2020\|EPI_ISL_680191\|202 | ···················· | ···················· | ···················· | ···················· | ···················· | ···················· |
| U | 7266 | hCoV-19/England/SHEF-D1ADD/2020\|EPI_ISL_680192\|202 | ···················· | ···················· | ···················· | ···················· | ···················· | ···················· |
| U | 7267 | hCoV-19/England/SHEF-D2043/2020\|EPI_ISL_680193\|202 | ···················· | ···················· | ···················· | ···················· | ···················· | ···················· |
| U | 7268 | hCoV-19/England/SHEF-D2FBF/2020\|EPI_ISL_680196\|202 | ···················· | ···················· | ···················· | ···················· | ·····T·············· | ···················· |
| U | 7269 | hCoV-19/Northern Ireland/NIRE-1094A0/2020\|EPI_ISL_ | ···················· | ···················· | ···················· | ···················· | ···················· | ···················· |
| U | 7270 | hCoV-19/Northern Ireland/NIRE-109525/2020\|EPI_ISL_ | ···················· | ···················· | ···················· | ···················· | ···················· | ···················· |
| U | 7271 | hCoV-19/Northern Ireland/NIRE-109604/2020\|EPI_ISL_ | ···················· | ···················· | ···················· | ···················· | ···················· | ···················· |
| U | 7272 | hCoV-19/Northern Ireland/NIRE-109613/2020\|EPI_ISL_ | ···················· | ···················· | ···················· | ···················· | ···················· | ···················· |
| U | 7273 | hCoV-19/Northern Ireland/NIRE-109640/2020\|EPI_ISL_ | ···················· | ···················· | ···················· | ···················· | ···················· | ···················· |
| U | 7274 | hCoV-19/Northern Ireland/NIRE-10966E/2020\|EPI_ISL_ | ···················· | ···················· | ···················· | ···················· | ···················· | ···················· |

| | | | | | | | | | |
|---|---|---|---|---|---|---|---|---|---|
| U | 7275 | hCoV-19/Northern Ireland/NIRE-10968C/2020\|EPI_ISL_ | ···················· | ···················· | ···················· | ···················· | ···················· | ···················· | |
| U | 7276 | hCoV-19/Northern Ireland/NIRE-10969B/2020\|EPI_ISL_ | ··················T· | ···················· | ···················· | ···················· | ···················· | ···················· | 1 |
| U | 7277 | hCoV-19/Northern Ireland/NIRE-1096AA/2020\|EPI_ISL_ | ···················· | ···················· | ···················· | ···················· | ···················· | ···················· | |
| U | 7278 | hCoV-19/Northern Ireland/NIRE-1096E6/2020\|EPI_ISL_ | ···················· | ···················· | ···················· | ···················· | ···················· | ···················· | |
| U | 7279 | hCoV-19/Northern Ireland/NIRE-109710/2020\|EPI_ISL_ | ···················· | ···················· | ···················· | ···················· | ···················· | ···················· | |
| U | 7280 | hCoV-19/Northern Ireland/NIRE-10973E/2020\|EPI_ISL_ | ···················· | ···················· | ···················· | ···················· | ···················· | ···················· | |
| U | 7281 | hCoV-19/Northern Ireland/NIRE-1097B6/2020\|EPI_ISL_ | ···················· | ···················· | ···················· | ···················· | ···················· | ···················· | |
| U | 7282 | hCoV-19/Northern Ireland/NIRE-1097F2/2020\|EPI_ISL_ | ···················· | ···················· | ···················· | ···················· | ···················· | ···················· | |
| U | 7283 | hCoV-19/Northern Ireland/NIRE-232E36/2020\|EPI_ISL_ | ···················· | ···················· | ···················· | ···················· | ···················· | ···················· | |
| U | 7284 | hCoV-19/England/MILK-BABE5D/2020\|EPI_ISL_681447\|20 | ···················· | ···················· | ···················· | ···················· | ···················· | ···················· | |
| U | 7285 | hCoV-19/England/MILK-BAABDF/2020\|EPI_ISL_681448\|20 | ···················· | ·A·················· | ···················· | ···················· | ···················· | ··A················· | |
| U | 7286 | hCoV-19/England/MILK-BACDE6/2020\|EPI_ISL_681449\|20 | ···················· | ···················· | ···················· | ···················· | ···················· | ···················· | |
| U | 7287 | hCoV-19/England/MILK-BAA89C/2020\|EPI_ISL_681450\|20 | ···················· | ···················· | ···················· | ···················· | ···················· | ···················· | |
| U | 7288 | hCoV-19/England/MILK-BABE99/2020\|EPI_ISL_681451\|20 | ···················· | ···················· | ···················· | ···················· | ···················· | ···················· | |
| U | 7289 | hCoV-19/England/MILK-BA9F3E/2020\|EPI_ISL_681452\|20 | ···················· | ···················· | ···················· | ···················· | ·····T·············· | ···················· | |
| U | 7290 | hCoV-19/England/MILK-BA9FA7/2020\|EPI_ISL_681454\|20 | ···················· | ···················· | ···················· | ···················· | ···················· | ···················· | |
| U | 7291 | hCoV-19/England/MILK-BAA36E/2020\|EPI_ISL_681455\|20 | ···················· | ···················· | ···················· | ···················· | ···················· | ···················· | |
| U | 7292 | hCoV-19/England/MILK-BAB039/2020\|EPI_ISL_681456\|20 | ···················· | ···················· | ···················· | ···················· | ·····T·············· | ···················· | |
| U | 7293 | hCoV-19/England/MILK-BAB01B/2020\|EPI_ISL_681457\|20 | ···················· | ···················· | ···················· | ···················· | ···················· | ···················· | |
| U | 7294 | hCoV-19/England/MILK-BABC44/2020\|EPI_ISL_681460\|20 | ···················· | ···················· | ···················· | ···················· | ···················· | ···················· | |
| U | 7295 | hCoV-19/England/MILK-BAA146/2020\|EPI_ISL_681461\|20 | ···················· | ···················· | ···················· | ···················· | ···················· | ···················· | |
| U | 7296 | hCoV-19/England/MILK-BABB74/2020\|EPI_ISL_681462\|20 | ···················· | ···················· | ···················· | ···················· | ···················· | ···················· | |
| U | 7297 | hCoV-19/England/MILK-BAAB84/2020\|EPI_ISL_681467\|20 | ···················· | ···················· | ···················· | ···················· | ···················· | ···················· | |
| U | 7298 | hCoV-19/England/MILK-BAA225/2020\|EPI_ISL_681469\|20 | ···················· | ·A·················· | ···················· | ···················· | ···················· | ··A················· | |
| U | 7299 | hCoV-19/England/MILK-BACD40/2020\|EPI_ISL_681470\|20 | ···················· | ···················· | ···················· | ···················· | ···················· | ···················· | |
| U | 7300 | hCoV-19/England/MILK-BAB479/2020\|EPI_ISL_681471\|20 | ···················· | ···················· | ···················· | ···················· | ···················· | ···················· | |
| U | 7301 | hCoV-19/England/MILK-BA99B8/2020\|EPI_ISL_681474\|20 | ···················· | ···················· | ···················· | ···················· | ···················· | ···················· | |
| U | 7302 | hCoV-19/England/MILK-BAAC36/2020\|EPI_ISL_681475\|20 | ···················· | ···················· | ···················· | ···················· | ···················· | ···················· | |
| U | 7303 | hCoV-19/England/MILK-BAB46A/2020\|EPI_ISL_681476\|20 | ···················· | ···················· | ···················· | ···················· | ···················· | ···················· | |
| U | 7304 | hCoV-19/England/MILK-BAB1AF/2020\|EPI_ISL_681478\|20 | ···················· | ···················· | ···················· | ···················· | ···················· | ···················· | |
| U | 7305 | hCoV-19/England/MILK-BAB998/2020\|EPI_ISL_681479\|20 | ···················· | ···················· | ···················· | ···················· | ···················· | ···················· | |
| U | 7306 | hCoV-19/England/MILK-BAB646/2020\|EPI_ISL_681480\|20 | ···················· | ···················· | ···················· | ···················· | ···················· | ···················· | |
| U | 7307 | hCoV-19/England/MILK-BAB50D/2020\|EPI_ISL_681481\|20 | ···················· | ···················· | ···················· | ···················· | ···················· | ···················· | |
| U | 7308 | hCoV-19/England/MILK-BAA920/2020\|EPI_ISL_681484\|20 | ···················· | ···················· | ···················· | ···················· | ···················· | ···················· | |
| U | 7309 | hCoV-19/England/MILK-BAB682/2020\|EPI_ISL_681485\|20 | ···················· | ···················· | ···················· | ···················· | ···················· | ···················· | |
| U | 7310 | hCoV-19/England/MILK-BAA7AE/2020\|EPI_ISL_681487\|20 | ···················· | ···················· | ···················· | ···················· | ···················· | ···················· | |
| U | 7311 | hCoV-19/England/MILK-BABC62/2020\|EPI_ISL_681488\|20 | ···················· | ···················· | ···················· | ···················· | ···················· | ···················· | |
| U | 7312 | hCoV-19/England/MILK-BAB242/2020\|EPI_ISL_681489\|20 | ···················· | ···················· | ···················· | ···················· | ···················· | ···················· | |
| U | 7313 | hCoV-19/England/MILK-BAAF97/2020\|EPI_ISL_681490\|20 | ···················· | ···················· | ···················· | ···················· | ···················· | ···················· | |
| U | 7314 | hCoV-19/England/MILK-BAC5FD/2020\|EPI_ISL_681492\|20 | ···················· | ·A·················· | ···················· | ···················· | ···················· | ···················· | |
| U | 7315 | hCoV-19/England/MILK-BAAE21/2020\|EPI_ISL_681493\|20 | ···················· | ···················· | ···················· | ···················· | ···················· | ···················· | |
| U | 7316 | hCoV-19/England/MILK-BAB048/2020\|EPI_ISL_681494\|20 | ···················· | ···················· | ···················· | ···················· | ···················· | ···················· | |
| U | 7317 | hCoV-19/England/MILK-BAB9B6/2020\|EPI_ISL_681495\|20 | ···················· | ···················· | ···················· | ···················· | ···················· | ···················· | |
| U | 7318 | hCoV-19/England/MILK-BAB260/2020\|EPI_ISL_681496\|20 | ···················· | ···················· | ···················· | ···················· | ···················· | ···················· | |
| U | 7319 | hCoV-19/England/MILK-BAA823/2020\|EPI_ISL_681497\|20 | ···················· | ···················· | ···················· | ···················· | ···················· | ···················· | |
| U | 7320 | hCoV-19/England/MILK-BAB5C1/2020\|EPI_ISL_681498\|20 | ···················· | ···················· | ···················· | ···················· | ···················· | ···················· | |
| U | 7321 | hCoV-19/England/MILK-BAAC27/2020\|EPI_ISL_681499\|20 | ···················· | ···················· | ···················· | ···················· | ···················· | ···················· | |
| U | 7322 | hCoV-19/England/MILK-BAAA96/2020\|EPI_ISL_681500\|20 | ···················· | ···················· | ···················· | ···················· | ···················· | ···················· | |
| U | 7323 | hCoV-19/England/MILK-BAC645/2020\|EPI_ISL_681501\|20 | ···················· | ···················· | ···················· | ···················· | ···················· | ···················· | |
| U | 7324 | hCoV-19/England/MILK-BAB9A7/2020\|EPI_ISL_681502\|20 | ···················· | ···················· | ···················· | ···················· | ···················· | ···················· | |
| U | 7325 | hCoV-19/England/MILK-BA9EAA/2020\|EPI_ISL_681503\|20 | ···················· | ···················· | ···················· | ···················· | ···················· | ···················· | |
| U | 7326 | hCoV-19/England/MILK-BABBFC/2020\|EPI_ISL_681504\|20 | ···················· | ···················· | ···················· | ···················· | ···················· | ···················· | |
| U | 7327 | hCoV-19/England/MILK-BAB2AC/2020\|EPI_ISL_681506\|20 | ···················· | ···················· | ···················· | ···················· | ···················· | ···················· | |
| U | 7328 | hCoV-19/England/MILK-BAA3AA/2020\|EPI_ISL_681507\|20 | ···················· | ···················· | ···················· | ···················· | ···················· | ···················· | |
| U | 7329 | hCoV-19/England/MILK-BAB79E/2020\|EPI_ISL_681508\|20 | ···················· | ···················· | ···················· | ···················· | ···················· | ···················· | |
| U | 7330 | hCoV-19/England/MILK-BA9B85/2020\|EPI_ISL_681509\|20 | ···················· | ···················· | ···················· | ···················· | ···················· | ···················· | |
| U | 7331 | hCoV-19/England/MILK-BAA94E/2020\|EPI_ISL_681510\|20 | ···················· | ···················· | ···················· | ···················· | ···················· | ···················· | |
| U | 7332 | hCoV-19/England/MILK-BAC681/2020\|EPI_ISL_681511\|20 | ···················· | ···················· | ···················· | ···················· | ·····T·············· | ···················· | |
| U | 7333 | hCoV-19/England/MILK-BA9AC4/2020\|EPI_ISL_681512\|20 | ···················· | ···················· | ···················· | ···················· | ···················· | ···················· | |
| U | 7334 | hCoV-19/England/MILK-BAB02A/2020\|EPI_ISL_681515\|20 | ···················· | ···················· | ···················· | ···················· | ···················· | ··A················· | |
| U | 7335 | hCoV-19/England/MILK-BACDB9/2020\|EPI_ISL_681516\|20 | ···················· | ···················· | ···················· | ···················· | ···················· | ···················· | |
| U | 7336 | hCoV-19/England/MILK-BAA5A4/2020\|EPI_ISL_681517\|20 | ···················· | ···················· | ···················· | ···················· | ···················· | ···················· | |
| U | 7337 | hCoV-19/England/MILK-BA9F01/2020\|EPI_ISL_681518\|20 | ···················· | ···················· | ···················· | ···················· | ···················· | ···················· | |
| U | 7338 | hCoV-19/England/MILK-BAAA87/2020\|EPI_ISL_681519\|20 | ···················· | ···················· | ···················· | ···················· | ···················· | ···················· | |
| U | 7339 | hCoV-19/England/MILK-BAAB66/2020\|EPI_ISL_681521\|20 | ···················· | ···················· | ···················· | ···················· | ···················· | ···················· | |
| U | 7340 | hCoV-19/England/MILK-BAB7F8/2020\|EPI_ISL_681522\|20 | ···················· | ···················· | ···················· | ···················· | ···················· | ···················· | |
| U | 7341 | hCoV-19/England/MILK-BAA3D7/2020\|EPI_ISL_681524\|20 | ···················· | ···················· | ···················· | ···················· | ···················· | ···················· | |
| U | 7342 | hCoV-19/England/MILK-BAA9D5/2020\|EPI_ISL_681526\|20 | ···················· | ···················· | ···················· | ···················· | ···················· | ···················· | |
| U | 7343 | hCoV-19/England/MILK-BAA5C2/2020\|EPI_ISL_681527\|20 | ···················· | ···················· | ···················· | ···················· | ···················· | ···················· | |
| U | 7344 | hCoV-19/England/MILK-BABB65/2020\|EPI_ISL_681530\|20 | ···················· | ···················· | ···················· | ···················· | ·····T·············· | ···················· | |
| U | 7345 | hCoV-19/England/MILK-BAAA2D/2020\|EPI_ISL_681531\|20 | ···················· | ···················· | ···················· | ···················· | ···················· | ···················· | |
| U | 7346 | hCoV-19/England/MILK-BAA3C8/2020\|EPI_ISL_681532\|20 | ···················· | ···················· | ···················· | ···················· | ···················· | ···················· | |
| U | 7347 | hCoV-19/England/MILK-BA99A9/2020\|EPI_ISL_681534\|20 | ···················· | ···················· | ···················· | ···················· | ···················· | ···················· | |
| U | 7348 | hCoV-19/England/MILK-BAB057/2020\|EPI_ISL_681535\|20 | ···················· | ···················· | ···················· | ···················· | ···················· | ···················· | |
| U | 7349 | hCoV-19/England/MILK-BAB4E2/2020\|EPI_ISL_681536\|20 | ···················· | ···················· | ···················· | ···················· | ···················· | ···················· | |
| U | 7350 | hCoV-19/England/MILK-BABE20/2020\|EPI_ISL_681537\|20 | ···················· | ···················· | ···················· | ···················· | ···················· | ···················· | |
| U | 7351 | hCoV-19/England/MILK-BAB9E3/2020\|EPI_ISL_681538\|20 | ···················· | ···················· | ···················· | ···················· | ···················· | ···················· | |
| U | 7352 | hCoV-19/England/MILK-BAA173/2020\|EPI_ISL_681540\|20 | ···················· | ·A·················· | ···················· | ···················· | ···················· | ··A················· | |
| U | 7353 | hCoV-19/England/MILK-BAB94D/2020\|EPI_ISL_681541\|20 | ···················· | ···················· | ···················· | ···················· | ···················· | ···················· | |
| U | 7354 | hCoV-19/England/MILK-BA997C/2020\|EPI_ISL_681542\|20 | ···················· | ···················· | ···················· | ···················· | ···················· | ···················· | |

```
U 7355 hCoV-19/England/MILK-BAA841/2020|EPI_ISL_681545|20 ···················· ···················· ····················· ···················· ···················· ····················
U 7356 hCoV-19/England/MILK-BAAB57/2020|EPI_ISL_681546|20 ···················· ···················· ····················· ···················· ···················· ····················
U 7357 hCoV-19/England/MILK-BAC654/2020|EPI_ISL_681548|20 ···················· ···················· ····················· ···················· ···················· ····················
U 7358 hCoV-19/England/MILK-BA9B94/2020|EPI_ISL_681549|20 ···················· ···················· ····················· ···················· ···················· ····················
U 7359 hCoV-19/England/MILK-BAAA1E/2020|EPI_ISL_681550|20 ···················· ···················· ····················· ···················· ······T············· ····················
U 7360 hCoV-19/England/MILK-BA9E9B/2020|EPI_ISL_681552|20 ···················· ···················· ····················· ···················· ···················· ····················
U 7361 hCoV-19/England/MILK-BABB83/2020|EPI_ISL_681553|20 ···················· ···················· ····················· ···················· ···················· ····················
U 7362 hCoV-19/England/MILK-BAABB1/2020|EPI_ISL_681554|20 ···················· ···················· ····················· ···················· ···················· ····················
U 7363 hCoV-19/Denmark/DCGC-12591/2020|EPI_ISL_682772|202 ···················· ···················· ····················· ···················· ···················· ····················
U 7364 hCoV-19/Denmark/DCGC-12601/2020|EPI_ISL_682782|202 ···················· ···················· ····················· ···················· ···················· ····················
U 7365 hCoV-19/Denmark/DCGC-12603/2020|EPI_ISL_682784|202 ···················· ·A·················· ····················· ···················· ···················· ····················
U 7366 hCoV-19/Denmark/DCGC-12604/2020|EPI_ISL_682785|202 ···················· ···················· ····················· ···················· ···················· ····················
U 7367 hCoV-19/Denmark/DCGC-12605/2020|EPI_ISL_682786|202 ···················· ···················· ····················· ···················· ···················· ····················
U 7368 hCoV-19/Denmark/DCGC-12606/2020|EPI_ISL_682787|202 ···················· ···················· ····················· ···················· ···················· ····················
U 7369 hCoV-19/Denmark/DCGC-12607/2020|EPI_ISL_682788|202 ···················· ···················· ····················· ···················· ···················· ····················
U 7370 hCoV-19/Denmark/DCGC-12608/2020|EPI_ISL_682789|202 ···················· ···················· ····················· ···················· ···················· ····················
U 7371 hCoV-19/Denmark/DCGC-12611/2020|EPI_ISL_682792|202 ···················· ···················· ····················· ···················· ···················· ····················
U 7372 hCoV-19/Denmark/DCGC-12612/2020|EPI_ISL_682793|202 ···················· ·A·················· ····················· ···················· ···················· ···A················
U 7373 hCoV-19/Denmark/DCGC-12614/2020|EPI_ISL_682795|202 ···················· ···················· ····················· ···················· ···················· ····················
U 7374 hCoV-19/Denmark/DCGC-12615/2020|EPI_ISL_682796|202 ···················· ···················· ····················· ···················· ···················· ····················
U 7375 hCoV-19/Denmark/DCGC-12617/2020|EPI_ISL_682798|202 ···················· ···················· ····················· ···················· ···················· ····················
U 7376 hCoV-19/Denmark/DCGC-12629/2020|EPI_ISL_682810|202 ···················· ···················· ····················· ···················· ···················· ····················
U 7377 hCoV-19/Denmark/DCGC-12630/2020|EPI_ISL_682811|202 ···················· ···················· ····················· ···················· ···················· ····················
U 7378 hCoV-19/Denmark/DCGC-12631/2020|EPI_ISL_682812|202 ···················· ···················· ····················· ···················· ···················· ····················
U 7379 hCoV-19/Denmark/DCGC-12632/2020|EPI_ISL_682813|202 ···················· ···················· ····················· ···················· ···················· ····················
U 7380 hCoV-19/Denmark/DCGC-12634/2020|EPI_ISL_682815|202 ···················· ···················· ····················· ···················· ···················· ····················
U 7381 hCoV-19/Denmark/DCGC-12636/2020|EPI_ISL_682817|202 ···················· ···················· ····················· ···················· ···················· ····················
U 7382 hCoV-19/Denmark/DCGC-12638/2020|EPI_ISL_682819|202 ···················· ···················· ····················· ···················· ···················· ····················
U 7383 hCoV-19/Denmark/DCGC-12639/2020|EPI_ISL_682820|202 ···················· ···················· ····················· ···················· ···················· ····················
U 7384 hCoV-19/Denmark/DCGC-12641/2020|EPI_ISL_682822|202 ···················· ···················· ····················· ···················· ···················· ····················
U 7385 hCoV-19/Denmark/DCGC-12642/2020|EPI_ISL_682823|202 ···················· ···················· ····················· ···················· ···················· ····················
U 7386 hCoV-19/Denmark/DCGC-12643/2020|EPI_ISL_682824|202 ···················· ···················· ····················· ···················· ···················· ····················
U 7387 hCoV-19/Denmark/DCGC-12646/2020|EPI_ISL_682827|202 ···················· ···················· ····················· ···················· ···················· ····················
U 7388 hCoV-19/Denmark/DCGC-12647/2020|EPI_ISL_682828|202 ···················· ···················· ····················· ···················· ···················· ····················
U 7389 hCoV-19/Denmark/DCGC-12650/2020|EPI_ISL_682831|202 ···················· ···················· ····················· ···················· ···················· ····················
U 7390 hCoV-19/Denmark/DCGC-12652/2020|EPI_ISL_682833|202 ···················· ···················· ····················· ···················· ···················· ····················
U 7391 hCoV-19/Denmark/DCGC-12654/2020|EPI_ISL_682835|202 ···················· ···················· ····················· ···················· ···················· ····················
U 7392 hCoV-19/Denmark/DCGC-12655/2020|EPI_ISL_682836|202 ···················· ···················· ····················· ···················· ···················· ····················
U 7393 hCoV-19/Denmark/DCGC-12656/2020|EPI_ISL_682837|202 ···················· ···················· ····················· ···················· ···················· ····················
U 7394 hCoV-19/Denmark/DCGC-12657/2020|EPI_ISL_682838|202 ···················· ···················· ····················· ···················· ···················· ····················
U 7395 hCoV-19/Denmark/DCGC-12659/2020|EPI_ISL_682840|202 ···················· ···················· ····················· ···················· ···················· ····················
U 7396 hCoV-19/Denmark/DCGC-12660/2020|EPI_ISL_682841|202 ···················· ···················· ·········A··········· ···················· ···················· ····················
U 7397 hCoV-19/Denmark/DCGC-12663/2020|EPI_ISL_682844|202 ···················· ···················· ····················· ···················· ···················· ····················
U 7398 hCoV-19/Denmark/DCGC-12664/2020|EPI_ISL_682845|202 ···················· ···················· ····················· ···················· ···················· ····················
U 7399 hCoV-19/Denmark/DCGC-12665/2020|EPI_ISL_682846|202 ···················· ·A·················· ····················· ···················· ···················· ····················
U 7400 hCoV-19/Denmark/DCGC-12667/2020|EPI_ISL_682848|202 ···················· ···················· ····················· ···················· ···················· ····················
U 7401 hCoV-19/Denmark/DCGC-12668/2020|EPI_ISL_682849|202 ···················· ···················· ····················· ···················· ···················· ····················
U 7402 hCoV-19/Denmark/DCGC-12669/2020|EPI_ISL_682850|202 ···················· ···················· ····················· ···················· ···················· ····················
U 7403 hCoV-19/Denmark/DCGC-12670/2020|EPI_ISL_682851|202 ···················· ···················· ····················· ···················· ···················· ····················
U 7404 hCoV-19/Denmark/DCGC-12673/2020|EPI_ISL_682854|202 ···················· ···················· ····················· ···················· ···················· ····················
U 7405 hCoV-19/Denmark/DCGC-12674/2020|EPI_ISL_682855|202 ···················· ···················· ····················· ···················· ···················· ····················
U 7406 hCoV-19/Denmark/DCGC-12675/2020|EPI_ISL_682856|202 ···················· ···················· ····················· ···················· ···················· ····················
U 7407 hCoV-19/Denmark/DCGC-12676/2020|EPI_ISL_682857|202 ···················· ···················· ····················· ···················· ···················· ····················
U 7408 hCoV-19/Denmark/DCGC-12677/2020|EPI_ISL_682858|202 ···················· ···················· ····················· ···················· ···················· ····················
U 7409 hCoV-19/Denmark/DCGC-12678/2020|EPI_ISL_682859|202 ···················· ···················· ····················· ···················· ···················· ····················
U 7410 hCoV-19/Denmark/DCGC-12681/2020|EPI_ISL_682862|202 ···················· ···················· ····················· ···················· ···················· ····················
U 7411 hCoV-19/Denmark/DCGC-12683/2020|EPI_ISL_682864|202 ···················· ····G··············· ····················· ···················· ···················· ····················
U 7412 hCoV-19/Denmark/DCGC-12685/2020|EPI_ISL_682866|202 ···················· ···················· ····················· ···················· ···················· ····················
U 7413 hCoV-19/Denmark/DCGC-12686/2020|EPI_ISL_682867|202 ···················· ···················· ····················· ···················· ···················· ····················
U 7414 hCoV-19/Denmark/DCGC-12687/2020|EPI_ISL_682868|202 ···················· ···················· ····················· ···················· ···················· ····················
U 7415 hCoV-19/Denmark/DCGC-12688/2020|EPI_ISL_682869|202 ···················· ···················· ····················· ···················· ···················· ····················
U 7416 hCoV-19/Denmark/DCGC-12689/2020|EPI_ISL_682870|202 ···················· ···················· ····················· ···················· ···················· ····················
U 7417 hCoV-19/Denmark/DCGC-12690/2020|EPI_ISL_682871|202 ···················· ···················· ····················· ···················· ···················· ····················
U 7418 hCoV-19/Denmark/DCGC-12693/2020|EPI_ISL_682874|202 ···················· ·A·················· ····················· ···················· ···················· ····················
U 7419 hCoV-19/Denmark/DCGC-12695/2020|EPI_ISL_682876|202 ···················· ···················· ····················· ···················· ···················· ····················
U 7420 hCoV-19/Denmark/DCGC-12697/2020|EPI_ISL_682878|202 ···················· ···················· ····················· ···················· ···················· ····················
U 7421 hCoV-19/Denmark/DCGC-12699/2020|EPI_ISL_682880|202 ···················· ···················· ····················· ···················· ···················· ····················
U 7422 hCoV-19/Denmark/DCGC-12700/2020|EPI_ISL_682881|202 ···················· ···················· ····················· ···················· ···················· ····················
U 7423 hCoV-19/Denmark/DCGC-12702/2020|EPI_ISL_682883|202 ···················· ···················· ····················· ···················· ···················· ····················
U 7424 hCoV-19/Denmark/DCGC-12704/2020|EPI_ISL_682885|202 ···················· ···················· ····················· ···················· ···················· ····················
U 7425 hCoV-19/England/CAMC-B0A074/2020|EPI_ISL_673002|20 ···················· ···················· ····················· ···················· ······T············· ····················
U 7426 hCoV-19/England/CAMC-B09F69/2020|EPI_ISL_673003|20 ···················· ···················· ····················· ···················· ···················· ····················
U 7427 hCoV-19/England/CAMC-B093E5/2020|EPI_ISL_673004|20 ···················· ···················· ····················· ···················· ···················· ····················
U 7428 hCoV-19/England/CAMC-B09181/2020|EPI_ISL_673005|20 ···················· ···················· ····················· ···················· ···················· ····················
U 7429 hCoV-19/England/CAMC-B094E2/2020|EPI_ISL_673006|20 ···················· ···················· ····················· ···················· ···················· ····················
U 7430 hCoV-19/England/CAMC-B09A1D/2020|EPI_ISL_673007|20 ···················· ···················· ····················· ···················· ···················· ····················
U 7431 hCoV-19/England/CAMC-B09C71/2020|EPI_ISL_673008|20 ···················· ···················· ····················· ···················· ···················· ····················
U 7432 hCoV-19/England/CAMC-B09039/2020|EPI_ISL_673009|20 ···················· ···················· ····················· ···················· ···················· ····················
U 7433 hCoV-19/England/CAMC-B094B5/2020|EPI_ISL_673010|20 ···················· ···················· ····················· ···················· ···················· ····················
U 7434 hCoV-19/England/CAMC-B08D60/2020|EPI_ISL_673011|20 ···················· ···················· ····················· ···················· ···················· ····················
```

U 7435 hCoV-19/England/CAMC-B09FA5/2020|EPI_ISL_673012|20 ···················· ···················· ····················· ···················· ······T············· ····················
U 7436 hCoV-19/England/CAMC-B099F2/2020|EPI_ISL_673013|20 ···················· ···················· ····················· ···················· ···················· ····················
U 7437 hCoV-19/England/CAMC-B095C1/2020|EPI_ISL_673014|20 ···················· ···················· ····················· ···················· ···················· ····················
U 7438 hCoV-19/England/CAMC-B09093/2020|EPI_ISL_673015|20 ···················· ···················· ····················· ···················· ···················· ····················
U 7439 hCoV-19/England/CAMC-B09479/2020|EPI_ISL_673016|20 ···················· ···················· ····················· ···················· ···················· ····················
U 7440 hCoV-19/England/CAMC-B09DC9/2020|EPI_ISL_673017|20 ···················· ···················· ····················· ···················· ···················· ····················
U 7441 hCoV-19/England/CAMC-B09576/2020|EPI_ISL_673019|20 ···················· ···················· ····················· ···················· ···················· ····················
U 7442 hCoV-19/England/CAMC-B08EC7/2020|EPI_ISL_673020|20 ···················· ···················· ····················· ···················· ···················· ····················
U 7443 hCoV-19/England/CAMC-B0A575/2020|EPI_ISL_673021|20 ···················· ···················· ····················· ···················· ···················· ····················
U 7444 hCoV-19/England/CAMC-B091DC/2020|EPI_ISL_673022|20 ···················· ···················· ················G···· ···················· ···················· ····················
U 7445 hCoV-19/England/CAMC-B08FE2/2020|EPI_ISL_673023|20 ···················· ···················· ····················· ···················· ···················· ····················
U 7446 hCoV-19/England/CAMC-B0A3C6/2020|EPI_ISL_673024|20 ···················· ···················· ····················· ···················· ···················· ····················
U 7447 hCoV-19/England/CAMC-B09EB7/2020|EPI_ISL_673025|20 ···················· ···················· ····················· ···················· ······T············· ····················
U 7448 hCoV-19/England/MILK-B8869A/2020|EPI_ISL_674269|20 ···················· ···················· ····················· ···················· ···················· ····················
U 7449 hCoV-19/England/ALDP-B84388/2020|EPI_ISL_674272|20 ···················· ···················· ····················· ···················· ···················· ····················
U 7450 hCoV-19/England/MILK-B8C177/2020|EPI_ISL_674274|20 ···················· ···················· ····················· ···················· ···················· ····················
U 7451 hCoV-19/England/MILK-B8872E/2020|EPI_ISL_674275|20 ···················· ···················· ····················· ···················· ···················· ····················
U 7452 hCoV-19/England/ALDP-B840DC/2020|EPI_ISL_674276|20 ···················· ···················· ····················· ···················· ···················· ····················
U 7453 hCoV-19/England/ALDP-B853A5/2020|EPI_ISL_674277|20 ···················· ···················· ····················· ···················· ···················· ····················
U 7454 hCoV-19/England/MILK-B8857F/2020|EPI_ISL_674279|20 ···················· ···················· ····················· ···················· ···················· ····················
U 7455 hCoV-19/England/MILK-B882F0/2020|EPI_ISL_674280|20 ···················· ···················· ····················· ···················· ······T············· ····················
U 7456 hCoV-19/England/ALDP-B84090/2020|EPI_ISL_674281|20 ···················· ···················· ····················· ···················· ···················· ····················
U 7457 hCoV-19/England/MILK-B88445/2020|EPI_ISL_674282|20 ···················· ···················· ····················· ···················· ···················· ····················
U 7458 hCoV-19/England/ALDP-B83D8B/2020|EPI_ISL_674283|20 ···················· ···················· ····················· ···················· ······K············· ····················
U 7459 hCoV-19/England/ALDP-B84FA2/2020|EPI_ISL_674284|20 ···················· ···················· ····················· ···················· ···················· ····················
U 7460 hCoV-19/England/ALDP-B85123/2020|EPI_ISL_674285|20 ···················· ···················· ····················· ···················· ···················· ····················
U 7461 hCoV-19/England/ALDP-B83DF4/2020|EPI_ISL_674286|20 ···················· ···················· ····················· ···················· ···················· ····················
U 7462 hCoV-19/England/ALDP-B84A47/2020|EPI_ISL_674287|20 ···················· ···················· ····················· ···················· ···················· ····················
U 7463 hCoV-19/England/MILK-B88120/2020|EPI_ISL_674289|20 ···················· ···················· ····················· ···················· ···················· ····················
U 7464 hCoV-19/England/MILK-B88287/2020|EPI_ISL_674290|20 ···················· ···················· ····················· ···················· ···················· ····················
U 7465 hCoV-19/England/MILK-B8C3F9/2020|EPI_ISL_674291|20 ···················· ···················· ····················· ···················· ···················· ····················
U 7466 hCoV-19/England/ALDP-B84EF0/2020|EPI_ISL_674292|20 ···················· ···················· ····················· ···················· ···················· ····················
U 7467 hCoV-19/England/MILK-B88278/2020|EPI_ISL_674293|20 ···················· ···················· ····················· ···················· ···················· ····················
U 7468 hCoV-19/England/ALDP-B8516F/2020|EPI_ISL_674294|20 ···················· ···················· ····················· ···················· ···················· ····················
U 7469 hCoV-19/England/ALDP-B84A92/2020|EPI_ISL_674295|20 ···················· ···················· ····················· ···················· ···················· ····················
U 7470 hCoV-19/England/ALDP-B83F67/2020|EPI_ISL_674296|20 ···················· ···················· ····················· ···················· ···················· ····················
U 7471 hCoV-19/England/ALDP-B83EF1/2020|EPI_ISL_674297|20 ···················· ···················· ····················· ···················· ···················· ····················
U 7472 hCoV-19/England/MILK-B8C1E0/2020|EPI_ISL_674298|20 ···················· ···················· ····················· ···················· ···················· ····················
U 7473 hCoV-19/England/ALDP-B8523F/2020|EPI_ISL_674300|20 ···················· ···················· ····················· ···················· ···················· ····················
U 7474 hCoV-19/England/ALDP-B8541B/2020|EPI_ISL_674301|20 ···················· ···················· ····················· ···················· ···················· ···A················
U 7475 hCoV-19/England/MILK-B8C2CF/2020|EPI_ISL_674302|20 ···················· ···················· ····················· ···················· ···················· ····················
U 7476 hCoV-19/England/MILK-B8C4C9/2020|EPI_ISL_674303|20 ···················· ···················· ····················· ···················· ···················· ····················
U 7477 hCoV-19/England/MILK-B8C450/2020|EPI_ISL_674304|20 ···················· ···················· ····················· ···················· ···················· ····················
U 7478 hCoV-19/England/ALDP-B83DA9/2020|EPI_ISL_674305|20 ···················· ···················· ····················· ···················· ···················· ····················
U 7479 hCoV-19/England/MILK-B88339/2020|EPI_ISL_674306|20 ···················· ···················· ····················· ···················· ···················· ····················
U 7480 hCoV-19/England/ALDP-B851C9/2020|EPI_ISL_674308|20 ···················· ···················· ····················· ···················· ···················· ····················
U 7481 hCoV-19/England/MILK-B88436/2020|EPI_ISL_674309|20 ···················· ···················· ····················· ···················· ···················· ····················
U 7482 hCoV-19/England/ALDP-B84BEA/2020|EPI_ISL_674310|20 ···················· ···················· ····················· ···················· ···················· ····················
U 7483 hCoV-19/England/ALDP-B839D2/2020|EPI_ISL_674311|20 ···················· ···················· ····················· ···················· ···················· ····················
U 7484 hCoV-19/England/ALDP-B851AB/2020|EPI_ISL_674313|20 ···················· ···················· ····················· ···················· ······T············· ····················
U 7485 hCoV-19/England/MILK-B8C274/2020|EPI_ISL_674314|20 ···················· ···················· ····················· ···················· ···················· ····················
U 7486 hCoV-19/England/MILK-B883A2/2020|EPI_ISL_674315|20 ···················· ···················· ····················· ···················· ···················· ····················
U 7487 hCoV-19/England/MILK-B8C0E3/2020|EPI_ISL_674316|20 ···················· ···················· ····················· ···················· ···················· ····················
U 7488 hCoV-19/England/ALDP-B83D7C/2020|EPI_ISL_674317|20 ···················· ···················· ····················· ···················· ···················· ····················
U 7489 hCoV-19/England/ALDP-B851E7/2020|EPI_ISL_674318|20 ···················· ·A·················· ····················· ···················· ···················· ···A················
U 7490 hCoV-19/England/ALDP-B84F48/2020|EPI_ISL_674319|20 ···················· ···················· ····················· ···················· ···················· ····················
U 7491 hCoV-19/England/MILK-B8C1FF/2020|EPI_ISL_674320|20 ···················· ···················· ····················· ···················· ···················· ····················
U 7492 hCoV-19/England/MILK-B8C3DB/2020|EPI_ISL_674321|20 ···················· ···················· ····················· ···················· ···················· ····················
U 7493 hCoV-19/England/ALDP-B83F1C/2020|EPI_ISL_674322|20 ···················· ···················· ····················· ···················· ···················· ····················
U 7494 hCoV-19/England/ALDP-B83F3A/2020|EPI_ISL_674323|20 ···················· ···················· ····················· ···················· ···················· ····················
U 7495 hCoV-19/England/MILK-B8873D/2020|EPI_ISL_674325|20 ···················· ···················· ····················· ···················· ···················· ····················
U 7496 hCoV-19/England/MILK-B8C098/2020|EPI_ISL_674326|20 ···················· ···················· ····················· ···················· ···················· ····················
U 7497 hCoV-19/England/ALDP-B84F57/2020|EPI_ISL_674328|20 ···················· ···················· ····················· ···················· ···················· ····················
U 7498 hCoV-19/England/ALDP-B83D03/2020|EPI_ISL_674329|20 ···················· ···················· ····················· ···················· ···················· ····················
U 7499 hCoV-19/England/MILK-B8858E/2020|EPI_ISL_674330|20 ···················· ···················· ····················· ···················· ···················· ····················
U 7500 hCoV-19/England/MILK-B8BFF6/2020|EPI_ISL_674331|20 ···················· ···················· ····················· ···················· ···················· ····················
U 7501 hCoV-19/England/ALDP-B84CBA/2020|EPI_ISL_674332|20 ···················· ···················· ····················· ···················· ···················· ····················
U 7502 hCoV-19/England/MILK-B884CD/2020|EPI_ISL_674333|20 ···················· ···················· ····················· ···················· ···················· ····················
U 7503 hCoV-19/England/MILK-B88515/2020|EPI_ISL_674335|20 ···················· ···················· ····················· ···················· ···················· ····················
U 7504 hCoV-19/England/MILK-B882B4/2020|EPI_ISL_674336|20 ···················· ·A·················· ····················· ···················· ···················· ····················
U 7505 hCoV-19/England/ALDP-B83E4C/2020|EPI_ISL_674337|20 ···················· ···················· ····················· ···················· ···················· ····················
U 7506 hCoV-19/England/ALDP-B843E2/2020|EPI_ISL_674339|20 ···················· ···················· ····················· ···················· ···················· ····················
U 7507 hCoV-19/England/ALDP-B8524E/2020|EPI_ISL_674340|20 ···················· ···················· ····················· ···················· ···················· ····················
U 7508 hCoV-19/England/ALDP-B84D02/2020|EPI_ISL_674341|20 ···················· ···················· ····················· ···················· ···················· ····················
U 7509 hCoV-19/England/MILK-B88357/2020|EPI_ISL_674342|20 ···················· ···················· ····················· ···················· ···················· ····················
U 7510 hCoV-19/England/ALDP-B85484/2020|EPI_ISL_674343|20 ···················· ···················· ····················· ···················· ······T············· ····················
U 7511 hCoV-19/England/MILK-B8BFD8/2020|EPI_ISL_674344|20 ···················· ···················· ····················· ···················· ···················· ····················
U 7512 hCoV-19/England/ALDP-B84F0C/2020|EPI_ISL_674345|20 ···················· ···················· ····················· ···················· ···················· ····················
U 7513 hCoV-19/England/ALDP-B84A65/2020|EPI_ISL_674346|20 ···················· ···················· ····················· ···················· ···················· ····················
U 7514 hCoV-19/England/ALDP-B83D4F/2020|EPI_ISL_674347|20 ···················· ···················· ····················· ···················· ···················· ····················

U 7515 hCoV-19/England/MILK-B8C362/2020|EPI_ISL_674349|20 ···················· ···················· ···················· ···················· ···················· ····················
U 7516 hCoV-19/England/ALDP-B84B35/2020|EPI_ISL_674351|20 ···················· ···················· ···················· ···················· ···················· ····················
U 7517 hCoV-19/England/ALDP-B853B4/2020|EPI_ISL_674352|20 ···················· ···················· ···················· ···················· ···················· ····················
U 7518 hCoV-19/England/ALDP-B83E1F/2020|EPI_ISL_674354|20 ···················· ···················· ···················· ···················· ···················· ····················
U 7519 hCoV-19/England/MILK-B8C414/2020|EPI_ISL_674355|20 ···················· ···················· ···················· ···················· ···················· ····················
U 7520 hCoV-19/England/MILK-B8C335/2020|EPI_ISL_674356|20 ···················· ···················· ···················· ···················· ···················· ····················
U 7521 hCoV-19/England/ALDP-B831E9/2020|EPI_ISL_675590|20 ···················· ···················· ···················· ···················· ···················· ····················
U 7522 hCoV-19/England/ALDP-B858F1/2020|EPI_ISL_675591|20 ···················· ···················· ···················· ···················· ···················· ····················
U 7523 hCoV-19/England/ALDP-B82997/2020|EPI_ISL_675592|20 ···················· ···················· ···················· ···················· ·····T·············· ····················
U 7524 hCoV-19/England/ALDP-B852C6/2020|EPI_ISL_675593|20 ···················· ···················· ···················· ···················· ···················· ····················
U 7525 hCoV-19/England/ALDP-B832C8/2020|EPI_ISL_675594|20 ···················· ···················· ···················· ···················· ···················· ····················
U 7526 hCoV-19/England/ALDP-B8583D/2020|EPI_ISL_675595|20 ···················· ···················· ···················· ···················· ···················· ····················
U 7527 hCoV-19/England/ALDP-B83529/2020|EPI_ISL_675597|20 ···················· ···················· ···················· ···················· ···················· ····················
U 7528 hCoV-19/England/ALDP-B85949/2020|EPI_ISL_675600|20 ···················· ···················· ···················· ···················· ···················· ····················
U 7529 hCoV-19/England/MILK-B8BF05/2020|EPI_ISL_675601|20 ···················· ···················· ···················· ···················· ···················· ····················
U 7530 hCoV-19/England/MILK-B89FCB/2020|EPI_ISL_675602|20 ···················· ·A·················· ···················· ···················· ···················· ····················
U 7531 hCoV-19/England/ALDP-B82AC1/2020|EPI_ISL_675603|20 ···················· ···················· ···················· ···················· ···················· ····················
U 7532 hCoV-19/England/ALDP-B858E2/2020|EPI_ISL_675604|20 ···················· ···················· ···················· ···················· ···················· ····················
U 7533 hCoV-19/England/ALDP-B92396/2020|EPI_ISL_675605|20 ···················· ···················· ···················· ···················· ···················· ····················
U 7534 hCoV-19/England/ALDP-B83301/2020|EPI_ISL_675607|20 ···················· ···················· ···················· ···················· ·····T·············· ····················
U 7535 hCoV-19/England/ALDP-B83055/2020|EPI_ISL_675608|20 ···················· ···················· ···················· ···················· ···················· ····················
U 7536 hCoV-19/England/ALDP-B8319E/2020|EPI_ISL_675609|20 ···················· ···················· ···················· ···················· ···················· ····················
U 7537 hCoV-19/England/ALDP-B85369/2020|EPI_ISL_675611|20 ···················· ···················· ···················· ···················· ···················· ····················
U 7538 hCoV-19/England/ALDP-B8326E/2020|EPI_ISL_675612|20 ···················· ···················· ···················· ···················· ···················· ····················
U 7539 hCoV-19/England/ALDP-B85A91/2020|EPI_ISL_675613|20 ···················· ···················· ···················· ···················· ···················· ····················
U 7540 hCoV-19/England/ALDP-B83B36/2020|EPI_ISL_675614|20 ···················· ···················· ···················· ···················· ···················· ····················
U 7541 hCoV-19/England/ALDP-B83B27/2020|EPI_ISL_675615|20 ···················· ···················· ···················· ···················· ···················· ····················
U 7542 hCoV-19/England/ALDP-B82A76/2020|EPI_ISL_675616|20 ···················· ···················· ···················· ···················· ·····T·············· ····················
U 7543 hCoV-19/England/ALDP-B85B8F/2020|EPI_ISL_675617|20 ···················· ···················· ···················· ···················· ···················· ····················
U 7544 hCoV-19/England/ALDP-B832D7/2020|EPI_ISL_675618|20 ···················· ···················· ···················· ···················· ···················· ····················
U 7545 hCoV-19/England/ALDP-B85ADD/2020|EPI_ISL_675619|20 ···················· ···················· ···················· ···················· ···················· ····················
U 7546 hCoV-19/England/MILK-B89F8F/2020|EPI_ISL_675621|20 ···················· ···················· ···················· ···················· ···················· ····················
U 7547 hCoV-19/England/ALDP-B83592/2020|EPI_ISL_675622|20 ···················· ···················· ···················· ···················· ·····T·············· ····················
U 7548 hCoV-19/England/ALDP-B8535A/2020|EPI_ISL_675623|20 ···················· ···················· ···················· ···················· ···················· ····················
U 7549 hCoV-19/England/ALDP-B8318F/2020|EPI_ISL_675624|20 ···················· ···················· ···················· ···················· ···················· ····················
U 7550 hCoV-19/England/ALDP-B82FB3/2020|EPI_ISL_675625|20 ···················· ···················· ···················· ···················· ···················· ····················
U 7551 hCoV-19/England/ALDP-B832F5/2020|EPI_ISL_675626|20 ···················· ···················· ···················· ···················· ···················· ····················
U 7552 hCoV-19/England/ALDP-B82A85/2020|EPI_ISL_675628|20 ···················· ·A·················· ···················· ···················· ···················· ····················
U 7553 hCoV-19/England/ALDP-B85A55/2020|EPI_ISL_675629|20 ···················· ···················· ···················· ···················· ···················· ····················
U 7554 hCoV-19/England/ALDP-B82A58/2020|EPI_ISL_675630|20 ···················· ···················· ···················· ···················· ···················· ····················
U 7555 hCoV-19/England/MILK-B89F9E/2020|EPI_ISL_675631|20 ···················· ···················· ···················· ···················· ···················· ····················
U 7556 hCoV-19/England/ALDP-B83161/2020|EPI_ISL_675632|20 ···················· ·A·················· ···················· ···················· ···················· ··A·················
U 7557 hCoV-19/England/ALDP-B9291C/2020|EPI_ISL_675633|20 ···················· ···················· ···················· ···················· ···················· ····················
U 7558 hCoV-19/England/MILK-B89FE9/2020|EPI_ISL_675634|20 ···················· ···················· ···················· ···················· ···················· ····················
U 7559 hCoV-19/England/ALDP-B82FC2/2020|EPI_ISL_675635|20 ···················· ···················· ···················· ···················· ···················· ····················
U 7560 hCoV-19/England/ALDP-B85A73/2020|EPI_ISL_675636|20 ···················· ···················· ···················· ···················· ···················· ····················
U 7561 hCoV-19/England/ALDP-B83073/2020|EPI_ISL_675637|20 ···················· ···················· ···················· ···················· ···················· ····················
U 7562 hCoV-19/England/ALDP-B83064/2020|EPI_ISL_675638|20 ···················· ···················· ···················· ···················· ···················· ····················
U 7563 hCoV-19/England/ALDP-B82EB6/2020|EPI_ISL_675639|20 ···················· ···················· ···················· ···················· ···················· ····················
U 7564 hCoV-19/England/ALDP-B83B81/2020|EPI_ISL_675640|20 ···················· ···················· ···················· ···················· ···················· ····················
U 7565 hCoV-19/England/ALDP-B83019/2020|EPI_ISL_675641|20 ···················· ···················· ···················· ···················· ·····T·············· ····················
U 7566 hCoV-19/England/ALDP-B854C0/2020|EPI_ISL_675642|20 ···················· ···················· ···················· ···················· ···················· ····················
U 7567 hCoV-19/England/ALDP-B92651/2020|EPI_ISL_675643|20 ···················· ···················· ···················· ···················· ···················· ····················
U 7568 hCoV-19/England/MILK-B89BF4/2020|EPI_ISL_675644|20 ···················· ···················· ···················· ···················· ·····T·············· ····················
U 7569 hCoV-19/England/ALDP-B8332F/2020|EPI_ISL_675645|20 ···················· ···················· ···················· ···················· ···················· ····················
U 7570 hCoV-19/England/ALDP-B83231/2020|EPI_ISL_675647|20 ···················· ···················· ···················· ···················· ···················· ····················
U 7571 hCoV-19/England/ALDP-B832AA/2020|EPI_ISL_675649|20 ···················· ···················· ···················· ···················· ···················· ····················
U 7572 hCoV-19/England/ALDP-B83B45/2020|EPI_ISL_675650|20 ···················· ···················· ···················· ···················· ···················· ····················
U 7573 hCoV-19/England/ALDP-B854DF/2020|EPI_ISL_675651|20 ···················· ···················· ···················· ···················· ···················· ····················
U 7574 hCoV-19/England/MILK-B89ECE/2020|EPI_ISL_675652|20 ···················· ···················· ···················· ···················· ···················· ····················
U 7575 hCoV-19/England/ALDP-B8350B/2020|EPI_ISL_675653|20 ···················· ···················· ···················· ···················· ···················· ····················
U 7576 hCoV-19/England/ALDP-B8300A/2020|EPI_ISL_675654|20 ···················· ···················· ···················· ···················· ···················· ····················
U 7577 hCoV-19/England/ALDP-B82FA4/2020|EPI_ISL_675655|20 ···················· ···················· ···················· ···················· ···················· ····················
U 7578 hCoV-19/England/ALDP-B927F4/2020|EPI_ISL_675656|20 ···················· ···················· ···················· ···················· ···················· ····················
U 7579 hCoV-19/England/ALDP-B83152/2020|EPI_ISL_675657|20 ···················· ···················· ···················· ···················· ···················· ··A·················
U 7580 hCoV-19/England/ALDP-B83556/2020|EPI_ISL_675658|20 ···················· ·A·················· ···················· ···················· ···················· ····················
U 7581 hCoV-19/England/ALDP-B82B46/2020|EPI_ISL_675659|20 ···················· ···················· ···················· ···················· ···················· ····················
U 7582 hCoV-19/England/MILK-B89DD0/2020|EPI_ISL_675660|20 ···················· ···················· ···················· ···················· ·····T·············· ····················
U 7583 hCoV-19/England/ALDP-B8530F/2020|EPI_ISL_675662|20 ···················· ···················· ···················· ···················· ···················· ····················
U 7584 hCoV-19/England/MILK-B89D76/2020|EPI_ISL_675663|20 ···················· ···················· ···················· ···················· ···················· ····················
U 7585 hCoV-19/England/ALDP-B82AA3/2020|EPI_ISL_675665|20 ···················· ·A·················· ···················· ···················· ···················· ····················
U 7586 hCoV-19/England/ALDP-B83046/2020|EPI_ISL_675666|20 ···················· ···················· ···················· ···················· ···················· ····················
U 7587 hCoV-19/England/MILK-B8BFBA/2020|EPI_ISL_675667|20 ···················· ···················· ···················· ···················· ···················· ····················
U 7588 hCoV-19/England/ALDP-B85967/2020|EPI_ISL_675668|20 ···················· ···················· ···················· ···················· ···················· ····················
U 7589 hCoV-19/England/ALDP-B83BBE/2020|EPI_ISL_675669|20 ···················· ···················· ···················· ···················· ···················· ····················
U 7590 hCoV-19/England/ALDP-B85A82/2020|EPI_ISL_675670|20 ···················· ···················· ···················· ···················· ···················· ····················
U 7591 hCoV-19/England/ALDP-B8590D/2020|EPI_ISL_675671|20 ···················· ···················· ···················· ···················· ···················· ····················
U 7592 hCoV-19/England/ALDP-B8593A/2020|EPI_ISL_675673|20 ···················· ···················· ···················· ···················· ···················· ····················
U 7593 hCoV-19/England/ALDP-B852D5/2020|EPI_ISL_675674|20 ···················· ···················· ···················· ···················· ···················· ····················
U 7594 hCoV-19/England/MILK-B89C6A/2020|EPI_ISL_675676|20 ···················· ···················· ···················· ···················· ···················· ····················

| | | | | | | | | |
|---|---|---|---|---|---|---|---|---|
| U | 7595 hCoV-19/England/ALDP-B82D22/2020\|EPI_ISL_675677\|20 | .................... | .................... | ..................... | .................... | .................... | .................... | |
| U | 7596 hCoV-19/England/ALDP-B8329B/2020\|EPI_ISL_675680\|20 | .................... | .................... | ..................... | .................... | .................... | .................... | |
| U | 7597 hCoV-19/England/ALDP-B92545/2020\|EPI_ISL_675681\|20 | .................... | .................... | ..................... | .................... | .................... | .................... | |
| U | 7598 hCoV-19/England/MILK-B89C88/2020\|EPI_ISL_675682\|20 | .................... | .................... | ..................... | .................... | .................... | .................... | |
| U | 7599 hCoV-19/England/ALDP-B85B07/2020\|EPI_ISL_675684\|20 | .................... | .................... | ..................... | .................... | .................... | .................... | |
| U | 7600 hCoV-19/England/ALDP-B92949/2020\|EPI_ISL_675685\|20 | .................... | .................... | ..................... | .................... | .................... | .................... | |
| U | 7601 hCoV-19/England/ALDP-B83B54/2020\|EPI_ISL_675686\|20 | .................... | .................... | ..................... | .................... | .................... | .................... | |
| U | 7602 hCoV-19/England/ALDP-B83BAF/2020\|EPI_ISL_675687\|20 | .................... | .................... | ..................... | .................... | ......T............. | .................... | |
| U | 7603 hCoV-19/USA/NY-Wadsworth-100017-01/2020\|EPI_ISL_67 | .................... | .................... | ..................... | .................... | .................... | .................... | |
| U | 7604 hCoV-19/USA/NY-Wadsworth-100184-01/2020\|EPI_ISL_67 | .................... | .................... | ..................... | .................... | .................... | .................... | |
| U | 7605 hCoV-19/USA/NY-Wadsworth-100463-01/2020\|EPI_ISL_67 | .................... | ..............C..... | ..................... | .................... | .................... | .................... | |
| U | 7606 hCoV-19/USA/NY-Wadsworth-101040-01/2020\|EPI_ISL_67 | .................... | .................... | ..................... | .................... | .................... | .................... | |
| U | 7607 hCoV-19/USA/NY-Wadsworth-101230-01/2020\|EPI_ISL_67 | .................... | .................... | ..................... | .................... | .................... | .................... | |
| U | 7608 hCoV-19/USA/NY-Wadsworth-102659-01/2020\|EPI_ISL_67 | .................... | .................... | ..................... | .................... | .................... | .................... | |
| U | 7609 hCoV-19/USA/NY-Wadsworth-102997-01/2020\|EPI_ISL_67 | .................... | .................... | ..................... | .................... | .................... | .................... | |
| U | 7610 hCoV-19/USA/NY-Wadsworth-103445-01/2020\|EPI_ISL_67 | .................... | .................... | ..................... | .................... | .................... | .................... | |
| U | 7611 hCoV-19/USA/NY-Wadsworth-103456-01/2020\|EPI_ISL_67 | .................... | .................... | ..................... | .................... | .................... | .................... | |
| U | 7612 hCoV-19/USA/NY-Wadsworth-103611-01/2020\|EPI_ISL_67 | .................... | .................... | ..................... | .................... | .................... | .................... | |
| U | 7614 hCoV-19/USA/NY-Wadsworth-105827-01/2020\|EPI_ISL_67 | .................... | .................... | ..................... | .................... | .................... | .................... | |
| U | 7615 hCoV-19/USA/NY-Wadsworth-106079-01/2020\|EPI_ISL_67 | .................... | .................... | ..................... | .................... | .................... | .................... | |
| U | 7616 hCoV-19/USA/NY-Wadsworth-106653-01/2020\|EPI_ISL_67 | .................... | .................... | ..................... | .................... | .................... | .................... | |
| U | 7617 hCoV-19/USA/NY-Wadsworth-106721-01/2020\|EPI_ISL_67 | .................... | .................... | ..................... | .................... | .................... | .................... | |
| U | 7618 hCoV-19/USA/NY-Wadsworth-109117-01/2020\|EPI_ISL_67 | .................... | .................... | ..................... | .................... | .................... | .................... | |
| U | 7619 hCoV-19/USA/NY-Wadsworth-109118-01/2020\|EPI_ISL_67 | .................... | .................... | ..................... | .................... | .................... | .................... | |
| U | 7620 hCoV-19/USA/NY-Wadsworth-109119-01/2020\|EPI_ISL_67 | .................... | .................... | ..................... | .................... | .................... | .................... | |
| U | 7621 hCoV-19/USA/NY-Wadsworth-109966-01/2020\|EPI_ISL_67 | .................... | .................... | ..................... | .................... | .................... | .................... | |
| U | 7622 hCoV-19/USA/NY-Wadsworth-110122-01/2020\|EPI_ISL_67 | .................... | .................... | ..................... | .................... | .................... | .................... | |
| U | 7623 hCoV-19/USA/NY-Wadsworth-111624-01/2020\|EPI_ISL_67 | .................... | .................... | ..................... | .................... | .................... | .................... | |
| U | 7624 hCoV-19/USA/NY-Wadsworth-114731-01/2020\|EPI_ISL_67 | .................... | .................... | ..................... | .................... | .................... | .................... | |
| U | 7625 hCoV-19/USA/NY-Wadsworth-114732-01/2020\|EPI_ISL_67 | .................... | .................... | ..................... | .................... | .................... | .................... | |
| U | 7626 hCoV-19/USA/NY-Wadsworth-116460-01/2020\|EPI_ISL_67 | .................... | .................... | ..................... | .................... | .................... | .................... | |
| U | 7628 hCoV-19/USA/NY-Wadsworth-118181-01/2020\|EPI_ISL_67 | .................... | .................... | ..................... | .................... | .................... | .................... | |
| U | 7629 hCoV-19/USA/NY-Wadsworth-122593-01/2020\|EPI_ISL_67 | .................... | .................... | ..................... | .................... | .................... | .................... | |
| U | 7630 hCoV-19/USA/NY-Wadsworth-125122-01/2020\|EPI_ISL_67 | .................... | .................... | ..................... | .................... | .................... | .................... | |
| U | 7631 hCoV-19/USA/NY-Wadsworth-126337-01/2020\|EPI_ISL_67 | .................... | .................... | ..................... | .................... | .................... | .................... | |
| U | 7632 hCoV-19/USA/NY-Wadsworth-142717-01/2020\|EPI_ISL_67 | .................... | .................... | ..................... | .................... | .................... | .................... | |
| U | 7633 hCoV-19/USA/NY-Wadsworth-142719-01/2020\|EPI_ISL_67 | .................... | .................... | ..................... | .................... | .................... | .................... | |
| U | 7634 hCoV-19/USA/NY-Wadsworth-142721-01/2020\|EPI_ISL_67 | .................... | .................... | ..................... | .................... | .................... | .................... | |
| U | 7635 hCoV-19/USA/NY-Wadsworth-142725-01/2020\|EPI_ISL_67 | .................... | .................... | ..................... | .................... | .................... | .................... | |
| U | 7636 hCoV-19/USA/NY-Wadsworth-142726-01/2020\|EPI_ISL_67 | .................... | .................... | ..................... | .................... | .................... | .................... | |
| U | 7637 hCoV-19/USA/NY-Wadsworth-142749-01/2020\|EPI_ISL_67 | .................... | .................... | ..................... | .................... | .................... | .................... | |
| U | 7638 hCoV-19/USA/NY-Wadsworth-142751-01/2020\|EPI_ISL_67 | .................... | .................... | ..................... | .................... | .................... | .................... | |
| U | 7639 hCoV-19/USA/NY-Wadsworth-142754-01/2020\|EPI_ISL_67 | .................... | .................... | ..................... | .................... | .................... | .................... | |
| U | 7640 hCoV-19/USA/NY-Wadsworth-142760-01/2020\|EPI_ISL_67 | .................... | .................... | ..................... | .................... | .................... | .................... | |
| U | 7641 hCoV-19/USA/NY-Wadsworth-142761-01/2020\|EPI_ISL_67 | .................... | .................... | ..................... | .................... | .................... | .................... | |
| U | 7642 hCoV-19/USA/NY-Wadsworth-142765-01/2020\|EPI_ISL_67 | .................... | .................... | ..................... | .................... | .................... | .................... | |
| U | 7643 hCoV-19/USA/NY-Wadsworth-142766-01/2020\|EPI_ISL_67 | .................... | .................... | ..................... | .................... | .................... | .................... | |
| U | 7644 hCoV-19/USA/NY-Wadsworth-142772-01/2020\|EPI_ISL_67 | .................... | .................... | ..................... | .................... | .................... | .................... | |
| U | 7645 hCoV-19/USA/NY-Wadsworth-142774-01/2020\|EPI_ISL_67 | .................... | .................... | ..................... | .................... | .................... | .................... | |
| U | 7646 hCoV-19/USA/NY-Wadsworth-142775-01/2020\|EPI_ISL_67 | .................... | .................... | ..................... | .................... | .................... | .................... | |
| U | 7647 hCoV-19/USA/NY-Wadsworth-142781-01/2020\|EPI_ISL_67 | .................... | .................... | ..................... | .................... | .................... | .................... | |
| U | 7648 hCoV-19/USA/NY-Wadsworth-142783-01/2020\|EPI_ISL_67 | .................... | .................... | ..................... | .................... | .................... | .................... | |
| U | 7649 hCoV-19/USA/NY-Wadsworth-142784-01/2020\|EPI_ISL_67 | .................... | .................... | ..................... | .................... | .................... | .................... | |
| U | 7650 hCoV-19/USA/NY-Wadsworth-142785-01/2020\|EPI_ISL_67 | .................... | .................... | ..................... | .................... | .................... | .................... | |
| U | 7651 hCoV-19/USA/NY-Wadsworth-143769-01/2020\|EPI_ISL_67 | .................... | ...................A | ..................... | .................... | .................... | .................... | 1 |
| U | 7652 hCoV-19/USA/NY-Wadsworth-144455-01/2020\|EPI_ISL_67 | .................... | .................... | ..................... | .................... | .................... | .................... | |
| U | 7653 hCoV-19/USA/NY-Wadsworth-144456-01/2020\|EPI_ISL_67 | .................... | .................... | ..................... | .................... | .................... | .................... | |
| U | 7654 hCoV-19/USA/NY-Wadsworth-146856-01/2020\|EPI_ISL_67 | .................... | .................... | ..................... | .................... | ......N............. | .................... | |
| U | 7655 hCoV-19/USA/NY-Wadsworth-152197-01/2020\|EPI_ISL_67 | .................... | ..............C..... | ..................... | .................... | .................... | .................... | |
| U | 7656 hCoV-19/USA/NY-Wadsworth-152368-01/2020\|EPI_ISL_67 | .................... | .................... | ..................... | .................... | .................... | .................... | |
| U | 7657 hCoV-19/USA/NY-Wadsworth-152382-01/2020\|EPI_ISL_67 | .................... | ..............C..... | ..................... | .................... | .................... | .................... | |
| U | 7658 hCoV-19/USA/NY-Wadsworth-152402-01/2020\|EPI_ISL_67 | .................... | ..............C..... | ..................... | .................... | .................... | .................... | |
| U | 7659 hCoV-19/USA/NY-Wadsworth-159719-01/2020\|EPI_ISL_67 | .................... | .................... | ..................... | .................... | .................... | .................... | |
| U | 7660 hCoV-19/USA/NY-Wadsworth-161631-01/2020\|EPI_ISL_67 | .................... | .A.................. | ..................... | .................... | .................... | .................... | |
| U | 7661 hCoV-19/USA/NY-Wadsworth-164501-01/2020\|EPI_ISL_67 | .................... | .................... | ..................... | .................... | .................... | .................... | |
| U | 7662 hCoV-19/USA/NY-Wadsworth-164567-01/2020\|EPI_ISL_67 | .................... | .................... | ..................... | .................... | .................... | .................... | |
| U | 7663 hCoV-19/USA/NY-Wadsworth-168120-01/2020\|EPI_ISL_67 | .................... | .................... | ..................... | .................... | .................... | .................... | |
| U | 7664 hCoV-19/USA/NY-Wadsworth-168141-01/2020\|EPI_ISL_67 | .................... | .................... | ..................... | .................... | .................... | .................... | |
| U | 7665 hCoV-19/USA/NY-Wadsworth-170604-01/2020\|EPI_ISL_67 | .................... | .................... | ..................... | .................... | .................... | .................... | |
| U | 7666 hCoV-19/USA/NY-Wadsworth-189267-01/2020\|EPI_ISL_67 | .................... | .................... | ..................... | .................... | .................... | .................... | |
| U | 7667 hCoV-19/USA/NY-Wadsworth-192863-01/2020\|EPI_ISL_67 | .................... | .................... | ..................... | .................... | .................... | .................... | |
| U | 7668 hCoV-19/USA/NY-Wadsworth-192864-01/2020\|EPI_ISL_67 | .................... | .................... | ..................... | .................... | .................... | .................... | |
| U | 7669 hCoV-19/USA/NY-Wadsworth-192865-01/2020\|EPI_ISL_67 | .................... | .................... | ..................... | .................... | .................... | .................... | |
| U | 7670 hCoV-19/USA/NY-Wadsworth-192866-01/2020\|EPI_ISL_67 | .................... | .................... | ..................... | .................... | .................... | .................... | |
| U | 7671 hCoV-19/USA/NY-Wadsworth-192867-01/2020\|EPI_ISL_67 | .................... | .................... | ..................... | .................... | .................... | .................... | |
| U | 7672 hCoV-19/England/SHEF-C150C/2020\|EPI_ISL_680115\|202 | .................... | .................... | ..................... | .................... | .................... | .................... | |
| U | 7673 hCoV-19/Northern Ireland/NIRE-1093C1/2020\|EPI_ISL_ | .................... | .................... | ..................... | .................... | .................... | .................... | |
| U | 7674 hCoV-19/Northern Ireland/NIRE-10940A/2020\|EPI_ISL_ | .................... | .................... | ..................... | .................... | .................... | .................... | |
| U | 7675 hCoV-19/England/MILK-BAB594/2020\|EPI_ISL_681442\|20 | .................... | .................... | ..................... | .................... | .................... | .................... | |
| U | 7676 hCoV-19/England/MILK-BAB1FA/2020\|EPI_ISL_681443\|20 | .................... | .................... | ..................... | .................... | .................... | .................... | |

| | | | | | | | |
|---|---|---|---|---|---|---|---|
| U | 7677 hCoV-19/England/MILK-BAAAE1/2020\|EPI_ISL_681444\|20 | ···················· | ···················· | ····················· | ···················· | ···················· | ···················· |
| U | 7678 hCoV-19/Denmark/DCGC-12592/2020\|EPI_ISL_682773\|202 | ···················· | ···················· | ····················· | ···················· | ···················· | ···················· |
| U | 7679 hCoV-19/Denmark/DCGC-12593/2020\|EPI_ISL_682774\|202 | ···················· | ···················· | ····················· | ···················· | ······T············· | ···················· |
| U | 7680 hCoV-19/Denmark/DCGC-12594/2020\|EPI_ISL_682775\|202 | ···················· | ···················· | ····················· | ···················· | ···················· | ···················· |
| U | 7681 hCoV-19/England/MILK-B8824B/2020\|EPI_ISL_674236\|20 | ···················· | ···················· | ····················· | ···················· | ···················· | ···················· |
| U | 7682 hCoV-19/England/MILK-B886B8/2020\|EPI_ISL_674238\|20 | ···················· | ···················· | ····················· | ···················· | ···················· | ···················· |
| U | 7683 hCoV-19/England/MILK-B886A9/2020\|EPI_ISL_674239\|20 | ···················· | ···················· | ····················· | ···················· | ···················· | ···················· |
| U | 7684 hCoV-19/England/ALDP-B8526C/2020\|EPI_ISL_674241\|20 | ···················· | ···················· | ····················· | ···················· | ···················· | ···················· |
| U | 7685 hCoV-19/England/ALDP-B83E79/2020\|EPI_ISL_674242\|20 | ···················· | ···················· | ····················· | ···················· | ···················· | ···················· |
| U | 7686 hCoV-19/England/MILK-B882D2/2020\|EPI_ISL_674244\|20 | ···················· | ···················· | ····················· | ···················· | ···················· | ···················· |
| U | 7687 hCoV-19/England/ALDP-B84F1B/2020\|EPI_ISL_674245\|20 | ···················· | ···················· | ····················· | ···················· | ···················· | ···················· |
| U | 7688 hCoV-19/England/MILK-B8C0A7/2020\|EPI_ISL_674246\|20 | ···················· | ···················· | ····················· | ···················· | ···················· | ···················· |
| U | 7689 hCoV-19/England/MILK-B8C3BD/2020\|EPI_ISL_674247\|20 | ···················· | ···················· | ····················· | ···················· | ···················· | ···················· |
| U | 7690 hCoV-19/England/MILK-B8C58A/2020\|EPI_ISL_674248\|20 | ···················· | ···················· | ····················· | ···················· | ···················· | ···················· |
| U | 7691 hCoV-19/England/ALDP-B843F1/2020\|EPI_ISL_674249\|20 | ···················· | ···················· | ····················· | ···················· | ···················· | ···················· |
| U | 7692 hCoV-19/England/MILK-B8C265/2020\|EPI_ISL_674250\|20 | ···················· | ···················· | ····················· | ···················· | ···················· | ···················· |
| U | 7693 hCoV-19/England/MILK-B8C2A1/2020\|EPI_ISL_674251\|20 | ···················· | ···················· | ····················· | ···················· | ···················· | ···················· |
| U | 7694 hCoV-19/England/MILK-B8C06B/2020\|EPI_ISL_674252\|20 | ···················· | ···················· | ····················· | ···················· | ···················· | ···················· |
| U | 7695 hCoV-19/England/ALDP-B849D1/2020\|EPI_ISL_674254\|20 | ···················· | ···················· | ····················· | ···················· | ···················· | ···················· |
| U | 7696 hCoV-19/England/ALDP-B843A6/2020\|EPI_ISL_674255\|20 | ···················· | ···················· | ····················· | ···················· | ···················· | ···················· |
| U | 7697 hCoV-19/England/MILK-B883EE/2020\|EPI_ISL_674257\|20 | ···················· | ···················· | ····················· | ···················· | ···················· | ···················· |
| U | 7698 hCoV-19/England/MILK-B88797/2020\|EPI_ISL_674258\|20 | ···················· | ···················· | ····················· | ···················· | ···················· | ···················· |
| U | 7699 hCoV-19/England/MILK-B88427/2020\|EPI_ISL_674259\|20 | ···················· | ···················· | ····················· | ···················· | ···················· | ···················· |
| U | 7700 hCoV-19/England/ALDP-B853C3/2020\|EPI_ISL_674260\|20 | ···················· | ···················· | ····················· | ···················· | ···················· | ···················· |
| U | 7701 hCoV-19/England/ALDP-B84ADE/2020\|EPI_ISL_674261\|20 | ···················· | ···················· | ····················· | ···················· | ···················· | ···················· |
| U | 7702 hCoV-19/England/MILK-B8C10E/2020\|EPI_ISL_674263\|20 | ···················· | ···················· | ····················· | ···················· | ···················· | ···················· |
| U | 7703 hCoV-19/England/ALDP-B83D9A/2020\|EPI_ISL_674264\|20 | ···················· | ···················· | ····················· | ···················· | ······T············· | ···················· |
| U | 7704 hCoV-19/England/MILK-B88788/2020\|EPI_ISL_674265\|20 | ···················· | ···················· | ····················· | ···················· | ···················· | ···················· |
| U | 7705 hCoV-19/England/ALDP-B85220/2020\|EPI_ISL_674267\|20 | ···················· | ···················· | ····················· | ···················· | ···················· | ···················· |
| U | 7706 hCoV-19/England/ALDP-B83082/2020\|EPI_ISL_675567\|20 | ···················· | ·A·················· | ····················· | ···················· | ···················· | ··A················· |
| U | 7707 hCoV-19/England/ALDP-B8333E/2020\|EPI_ISL_675568\|20 | ···················· | ···················· | ····················· | ···················· | ···················· | ···················· |
| U | 7708 hCoV-19/England/ALDP-B8328C/2020\|EPI_ISL_675570\|20 | ···················· | ···················· | ····················· | ···················· | ···················· | ···················· |
| U | 7709 hCoV-19/England/ALDP-B830CE/2020\|EPI_ISL_675571\|20 | ···················· | ···················· | ····················· | ···················· | ···················· | ···················· |
| U | 7710 hCoV-19/England/ALDP-B83BDC/2020\|EPI_ISL_675573\|20 | ···················· | ···················· | ····················· | ···················· | ···················· | ···················· |
| U | 7711 hCoV-19/England/ALDP-B82AFE/2020\|EPI_ISL_675574\|20 | ···················· | ···················· | ····················· | ···················· | ···················· | ···················· |
| U | 7712 hCoV-19/England/ALDP-B8533C/2020\|EPI_ISL_675575\|20 | ···················· | ···················· | ····················· | ···················· | ···················· | ···················· |
| U | 7713 hCoV-19/England/MILK-B89D94/2020\|EPI_ISL_675576\|20 | ···················· | ···················· | ····················· | ···················· | ···················· | ···················· |
| U | 7714 hCoV-19/England/MILK-B89E82/2020\|EPI_ISL_675578\|20 | ···················· | ···················· | ····················· | ···················· | ···················· | ···················· |
| U | 7715 hCoV-19/England/ALDP-B82E4D/2020\|EPI_ISL_675580\|20 | ···················· | ···················· | ····················· | ···················· | ···················· | ···················· |
| U | 7716 hCoV-19/England/ALDP-B83B90/2020\|EPI_ISL_675581\|20 | ···················· | ···················· | ····················· | ···················· | ···················· | ···················· |
| U | 7717 hCoV-19/England/ALDP-B82B37/2020\|EPI_ISL_675582\|20 | ···················· | ···················· | ····················· | ···················· | ···················· | ···················· |
| U | 7718 hCoV-19/England/ALDP-B8586A/2020\|EPI_ISL_675584\|20 | ···················· | ···················· | ····················· | ···················· | ···················· | ···················· |
| U | 7719 hCoV-19/England/ALDP-B85AEC/2020\|EPI_ISL_675585\|20 | ···················· | ···················· | ····················· | ···················· | ······T············· | ···················· |
| U | 7720 hCoV-19/England/ALDP-B8585B/2020\|EPI_ISL_675586\|20 | ···················· | ···················· | ····················· | ···················· | ···················· | ···················· |
| U | 7721 hCoV-19/England/ALDP-B858B5/2020\|EPI_ISL_675587\|20 | ···················· | ···················· | ····················· | ···················· | ···················· | ···················· |
| U | 7722 hCoV-19/England/ALDP-B85B61/2020\|EPI_ISL_675588\|20 | ···················· | ···················· | ····················· | ···················· | ···················· | ···················· |
| U | 7723 hCoV-19/England/ALDP-B92448/2020\|EPI_ISL_675589\|20 | ···················· | ···················· | ····················· | ···················· | ···················· | ···················· |
| U | 7724 hCoV-19/England/ALDP-B852F3/2020\|EPI_ISL_675598\|20 | ···················· | ···················· | ····················· | ···················· | ···················· | ···················· |
| U | 7725 hCoV-19/England/ALDP-B831F8/2020\|EPI_ISL_675599\|20 | ···················· | ···················· | ····················· | ···················· | ···················· | ···················· |
| U | 7726 hCoV-19/USA/NY-Wadsworth-76676-01/2020\|EPI_ISL_676 | ···················· | ···················· | ····················· | ···················· | ···················· | ···················· |
| U | 7727 hCoV-19/USA/NY-Wadsworth-76680-01/2020\|EPI_ISL_676 | ···················· | ···················· | ····················· | ···················· | ···················· | ···················· |
| U | 7728 hCoV-19/England/BIRM-6A927/2020\|EPI_ISL_679100\|202 | ···················· | ···················· | ····················· | ···················· | ···················· | ···················· |
| U | 7729 hCoV-19/England/BIRM-6A936/2020\|EPI_ISL_679101\|202 | ···················· | ···················· | ····················· | ···················· | ···················· | ···················· |
| U | 7730 hCoV-19/England/BIRM-6A945/2020\|EPI_ISL_679102\|202 | ···················· | ···················· | ····················· | ···················· | ···················· | ···················· |
| U | 7731 hCoV-19/England/QEUH-B2D13C/2020\|EPI_ISL_680100\|20 | ···················· | ···················· | ····················· | ···················· | ···················· | ···················· |
| U | 7732 hCoV-19/England/MILK-BA9B1C/2020\|EPI_ISL_681400\|20 | ···················· | ···················· | ····················· | ···················· | ···················· | ···················· |
| U | 7733 hCoV-19/England/MILK-BAA1DD/2020\|EPI_ISL_681401\|20 | ···················· | ···················· | ····················· | ···················· | ···················· | ···················· |
| U | 7734 hCoV-19/England/MILK-BA9BC1/2020\|EPI_ISL_681402\|20 | ···················· | ···················· | ····················· | ···················· | ···················· | ···················· |
| U | 7735 hCoV-19/England/MILK-BAAAC3/2020\|EPI_ISL_681403\|20 | ···················· | ···················· | ····················· | ···················· | ···················· | ···················· |
| U | 7736 hCoV-19/England/MILK-BAB5FE/2020\|EPI_ISL_681404\|20 | ···················· | ···················· | ····················· | ···················· | ···················· | ···················· |
| U | 7737 hCoV-19/England/MILK-BAAB39/2020\|EPI_ISL_681405\|20 | ···················· | ···················· | ····················· | ···················· | ···················· | ···················· |
| U | 7738 hCoV-19/England/MILK-BABE7B/2020\|EPI_ISL_681406\|20 | ···················· | ···················· | ····················· | ···················· | ······T············· | ···················· |
| U | 7739 hCoV-19/England/MILK-BAB549/2020\|EPI_ISL_681407\|20 | ···················· | ···················· | ····················· | ···················· | ···················· | ···················· |
| U | 7740 hCoV-19/England/MILK-BA9A88/2020\|EPI_ISL_681408\|20 | ···················· | ·A·················· | ····················· | ···················· | ···················· | ··A················· |
| U | 7741 hCoV-19/England/MILK-BACDC8/2020\|EPI_ISL_681409\|20 | ···················· | ···················· | ····················· | ···················· | ···················· | ··A················· |
| U | 7742 hCoV-19/England/MILK-BAAF88/2020\|EPI_ISL_681411\|20 | ···················· | ···················· | ····················· | ···················· | ···················· | ···················· |
| U | 7743 hCoV-19/England/MILK-BAA049/2020\|EPI_ISL_681412\|20 | ···················· | ···················· | ····················· | ···················· | ···················· | ···················· |
| U | 7744 hCoV-19/England/MILK-BAA87E/2020\|EPI_ISL_681414\|20 | ···················· | ···················· | ····················· | ···················· | ···················· | ···················· |
| U | 7745 hCoV-19/England/MILK-BAC663/2020\|EPI_ISL_681415\|20 | ···················· | ···················· | ····················· | ···················· | ···················· | ···················· |
| U | 7746 hCoV-19/England/MILK-BAB628/2020\|EPI_ISL_681416\|20 | ············T······· | ···················· | ····················· | ···················· | ···················· | ···················· |
| U | 7747 hCoV-19/England/MILK-BAA401/2020\|EPI_ISL_681417\|20 | ···················· | ···················· | ····················· | ···················· | ···················· | ···················· |
| U | 7748 hCoV-19/England/MILK-BAC6BE/2020\|EPI_ISL_681418\|20 | ···················· | ···················· | ····················· | ···················· | ···················· | ···················· |
| U | 7749 hCoV-19/England/MILK-BAA911/2020\|EPI_ISL_681419\|20 | ···················· | ·A·················· | ····················· | ···················· | ···················· | ···················· |
| U | 7750 hCoV-19/England/MILK-BA999A/2020\|EPI_ISL_681420\|20 | ···················· | ···················· | ····················· | ···················· | ···················· | ···················· |
| U | 7751 hCoV-19/England/MILK-BAC609/2020\|EPI_ISL_681421\|20 | ···················· | ·A·················· | ····················· | ···················· | ···················· | ···················· |
| U | 7752 hCoV-19/England/MILK-BAC6CD/2020\|EPI_ISL_681422\|20 | ···················· | ···················· | ····················· | ···················· | ···················· | ···················· |
| U | 7753 hCoV-19/England/MILK-BAB93E/2020\|EPI_ISL_681426\|20 | ···················· | ···················· | ····················· | ···················· | ···················· | ···················· |
| U | 7754 hCoV-19/England/MILK-BA9EC8/2020\|EPI_ISL_681427\|20 | ···················· | ···················· | ····················· | ···················· | ···················· | ···················· |
| U | 7755 hCoV-19/England/MILK-BA9E8C/2020\|EPI_ISL_681428\|20 | ···················· | ···················· | ····················· | ···················· | ······T············· | ···················· |
| U | 7756 hCoV-19/England/MILK-BAA805/2020\|EPI_ISL_681429\|20 | ···················· | ···················· | ····················· | ···················· | ···················· | ···················· |

| | | | | | | | | |
|---|---|---|---|---|---|---|---|---|
| U | 7757 | hCoV-19/England/MILK-BAA331/2020\|EPI_ISL_681430\|20 | .................... | .................... | .................... | .................... | .................... | .................... |
| U | 7758 | hCoV-19/England/MILK-BAB664/2020\|EPI_ISL_681431\|20 | .................... | .................... | .................... | .................... | .................... | .................... |
| U | 7759 | hCoV-19/England/MILK-BA9F10/2020\|EPI_ISL_681432\|20 | .................... | .................... | .................... | .................... | .................... | .................... |
| U | 7760 | hCoV-19/England/MILK-BAA7BD/2020\|EPI_ISL_681433\|20 | .................... | .................... | .................... | .................... | .................... | .................... |
| U | 7761 | hCoV-19/England/MILK-BAB585/2020\|EPI_ISL_681434\|20 | .................... | .................... | .................... | .................... | .................... | .................... |
| U | 7762 | hCoV-19/England/MILK-BABBDE/2020\|EPI_ISL_681437\|20 | .................... | .................... | .................... | .................... | .................... | .................... |
| U | 7763 | hCoV-19/England/MILK-BAB4F1/2020\|EPI_ISL_681438\|20 | .................... | .................... | .................... | .................... | .................... | .................... |
| U | 7764 | hCoV-19/England/MILK-BA9A79/2020\|EPI_ISL_681440\|20 | .................... | .................... | .................... | .................... | .................... | .................... |
| U | 7765 | hCoV-19/Denmark/DCGC-12521/2020\|EPI_ISL_682702\|202 | .................... | .................... | .................... | .................... | .................... | .................... |
| U | 7766 | hCoV-19/Denmark/DCGC-12522/2020\|EPI_ISL_682703\|202 | .................... | .................... | .................... | .................... | .................... | .................... |
| U | 7767 | hCoV-19/Denmark/DCGC-12524/2020\|EPI_ISL_682705\|202 | .................... | .A.................. | .................... | .................... | .................... | .................... |
| U | 7768 | hCoV-19/Denmark/DCGC-12525/2020\|EPI_ISL_682706\|202 | .................... | .................... | .................... | .................... | .................... | .................... |
| U | 7769 | hCoV-19/Denmark/DCGC-12526/2020\|EPI_ISL_682707\|202 | .................... | .................... | .................... | .................... | .................... | .................... |
| U | 7770 | hCoV-19/Denmark/DCGC-12527/2020\|EPI_ISL_682708\|202 | .................... | .................... | .................... | .................... | .................... | .................... |
| U | 7771 | hCoV-19/Denmark/DCGC-12528/2020\|EPI_ISL_682709\|202 | .................... | .................... | .................... | .................... | .................... | .................... |
| U | 7772 | hCoV-19/Denmark/DCGC-12529/2020\|EPI_ISL_682710\|202 | .................... | .................... | .................... | .................... | .................... | .................... |
| U | 7773 | hCoV-19/Denmark/DCGC-12530/2020\|EPI_ISL_682711\|202 | .................... | .................... | .................... | .................... | .................... | .................... |
| U | 7774 | hCoV-19/Denmark/DCGC-12531/2020\|EPI_ISL_682712\|202 | .................... | .................... | .................... | .................... | .................... | .................... |
| U | 7775 | hCoV-19/Denmark/DCGC-12532/2020\|EPI_ISL_682713\|202 | .................... | .................... | .................... | .................... | .................... | .................... |
| U | 7776 | hCoV-19/Denmark/DCGC-12536/2020\|EPI_ISL_682717\|202 | .................... | .................... | .................... | .................... | .................... | .................... |
| U | 7777 | hCoV-19/Denmark/DCGC-12537/2020\|EPI_ISL_682718\|202 | .................... | .................... | .................... | .................... | .................... | .................... |
| U | 7778 | hCoV-19/Denmark/DCGC-12538/2020\|EPI_ISL_682719\|202 | .................... | .................... | .................... | .................... | .................... | .................... |
| U | 7779 | hCoV-19/Denmark/DCGC-12539/2020\|EPI_ISL_682720\|202 | .................... | .................... | .................... | .................... | .................... | .................... |
| U | 7780 | hCoV-19/Denmark/DCGC-12540/2020\|EPI_ISL_682721\|202 | .................... | .................... | .................... | .................... | .................... | .................... |
| U | 7781 | hCoV-19/Denmark/DCGC-12541/2020\|EPI_ISL_682722\|202 | .................... | .................... | .................... | .................... | .....T.............. | .................... |
| U | 7782 | hCoV-19/Denmark/DCGC-12542/2020\|EPI_ISL_682723\|202 | .................... | .................... | .................... | .................... | .................... | .................... |
| U | 7783 | hCoV-19/Denmark/DCGC-12543/2020\|EPI_ISL_682724\|202 | .................... | .................... | .................... | .................... | .................... | .................... |
| U | 7784 | hCoV-19/Denmark/DCGC-12545/2020\|EPI_ISL_682726\|202 | .................... | .................... | .................... | .................... | .................... | .................... |
| U | 7785 | hCoV-19/Denmark/DCGC-12546/2020\|EPI_ISL_682727\|202 | .................... | .................... | .................... | .................... | .................... | .................... |
| U | 7786 | hCoV-19/Denmark/DCGC-12547/2020\|EPI_ISL_682728\|202 | .................... | .................... | .................... | .................... | .................... | .................... |
| U | 7787 | hCoV-19/Denmark/DCGC-12549/2020\|EPI_ISL_682730\|202 | .................... | .................... | .................... | .................... | .................... | .................... |
| U | 7788 | hCoV-19/Denmark/DCGC-12550/2020\|EPI_ISL_682731\|202 | .................... | .................... | .................... | .................... | .................... | .................... |
| U | 7789 | hCoV-19/Denmark/DCGC-12551/2020\|EPI_ISL_682732\|202 | .................... | .................... | .................... | .................... | .................... | .................... |
| U | 7790 | hCoV-19/Denmark/DCGC-12552/2020\|EPI_ISL_682733\|202 | .................... | .................... | .................... | .................... | .................... | .................... |
| U | 7791 | hCoV-19/Denmark/DCGC-12553/2020\|EPI_ISL_682734\|202 | .................... | .................... | .................... | .................... | .................... | .................... |
| U | 7792 | hCoV-19/Denmark/DCGC-12555/2020\|EPI_ISL_682736\|202 | .................... | .................... | .................... | .................... | .................... | .................... |
| U | 7793 | hCoV-19/Denmark/DCGC-12560/2020\|EPI_ISL_682741\|202 | .................... | .................... | .................... | .................... | .................... | .................... |
| U | 7794 | hCoV-19/Denmark/DCGC-12561/2020\|EPI_ISL_682742\|202 | .................... | .................... | .................... | .................... | .................... | .................... |
| U | 7795 | hCoV-19/Denmark/DCGC-12563/2020\|EPI_ISL_682744\|202 | .................... | .................... | .................... | .................... | .................... | .................... |
| U | 7796 | hCoV-19/Denmark/DCGC-12565/2020\|EPI_ISL_682746\|202 | .................... | ...A................ | .................... | .................... | .................... | .................... |
| U | 7797 | hCoV-19/Denmark/DCGC-12566/2020\|EPI_ISL_682747\|202 | .................... | .................... | .................... | .................... | .................... | .................... |
| U | 7798 | hCoV-19/Denmark/DCGC-12567/2020\|EPI_ISL_682748\|202 | .................... | .................... | .................... | .................... | .................... | .................... |
| U | 7799 | hCoV-19/Denmark/DCGC-12568/2020\|EPI_ISL_682749\|202 | .................... | .................... | .................... | .................... | .................... | .................... |
| U | 7800 | hCoV-19/Denmark/DCGC-12569/2020\|EPI_ISL_682750\|202 | .................... | .A.................. | .................... | .................... | .................... | .................... |
| U | 7801 | hCoV-19/Denmark/DCGC-12570/2020\|EPI_ISL_682751\|202 | .................... | .................... | .................... | .................... | .................... | .................... |
| U | 7802 | hCoV-19/Denmark/DCGC-12571/2020\|EPI_ISL_682752\|202 | .................... | .................... | .................... | .................... | .................... | .................... |
| U | 7803 | hCoV-19/Denmark/DCGC-12572/2020\|EPI_ISL_682753\|202 | .................... | .................... | .................... | .................... | .................... | .................... |
| U | 7804 | hCoV-19/Denmark/DCGC-12573/2020\|EPI_ISL_682754\|202 | .................... | .................... | .................... | .................... | .................... | .................... |
| U | 7805 | hCoV-19/Denmark/DCGC-12576/2020\|EPI_ISL_682757\|202 | .................... | .................... | .................... | .................... | .................... | .................... |
| U | 7806 | hCoV-19/Denmark/DCGC-12577/2020\|EPI_ISL_682758\|202 | .................... | .................... | .................... | .................... | .................... | .................... |
| U | 7807 | hCoV-19/Denmark/DCGC-12578/2020\|EPI_ISL_682759\|202 | .................... | .A.................. | .................... | .................... | .................... | .................... |
| U | 7808 | hCoV-19/Denmark/DCGC-12580/2020\|EPI_ISL_682761\|202 | .................... | .................... | .................... | .................... | .................... | .................... |
| U | 7809 | hCoV-19/Denmark/DCGC-12582/2020\|EPI_ISL_682763\|202 | .................... | .................... | .................... | .................... | .................... | .................... |
| U | 7810 | hCoV-19/Denmark/DCGC-12583/2020\|EPI_ISL_682764\|202 | .................... | .................... | .................... | .................... | .................... | .................... |
| U | 7811 | hCoV-19/Denmark/DCGC-12585/2020\|EPI_ISL_682766\|202 | .................... | .................... | .................... | .................... | .................... | .................... |
| U | 7812 | hCoV-19/Denmark/DCGC-12587/2020\|EPI_ISL_682768\|202 | .................... | .................... | .................... | .................... | .................... | .................... |
| U | 7813 | hCoV-19/Denmark/DCGC-12588/2020\|EPI_ISL_682769\|202 | .................... | .................... | .................... | .................... | .................... | .................... |
| V | 1 | hCoV-19/Japan/PG-0790/2020\|EPI_ISL_684400\|2020-04 | .................... | .................... | .................... | .................... | .................... | .................... |
| V | 2 | hCoV-19/Japan/PG-0791/2020\|EPI_ISL_684401\|2020-04 | .................... | .................... | .................... | .................... | .................... | .................... |
| V | 3 | hCoV-19/Japan/PG-0792/2020\|EPI_ISL_684402\|2020-04 | .................... | .................... | .................... | .................... | .................... | .................... |
| V | 4 | hCoV-19/Japan/PG-0793/2020\|EPI_ISL_684403\|2020-04 | .................... | .................... | .................... | .................... | .................... | .................... |
| V | 5 | hCoV-19/Japan/PG-0794/2020\|EPI_ISL_684404\|2020-04 | .................... | .................... | .................... | .................... | .................... | .................... |
| V | 6 | hCoV-19/Japan/PG-0795/2020\|EPI_ISL_684405\|2020-04 | .................... | .................... | .................... | .................... | .................... | .................... |
| V | 7 | hCoV-19/Japan/PG-0796/2020\|EPI_ISL_684406\|2020-04 | .................... | .................... | .................... | .................... | .................... | .................... |
| V | 8 | hCoV-19/Japan/PG-0797/2020\|EPI_ISL_684407\|2020-04 | .................... | .................... | .................... | .................... | .................... | .................... |
| V | 9 | hCoV-19/Japan/PG-0798/2020\|EPI_ISL_684408\|2020-04 | .................... | .................... | .................... | .................... | .................... | .................... |
| V | 10 | hCoV-19/Japan/PG-0799/2020\|EPI_ISL_684409\|2020-04 | .................... | .................... | .................... | .................... | .................... | .................... |
| V | 11 | hCoV-19/Japan/PG-0800/2020\|EPI_ISL_684410\|2020-04 | .................... | .................... | .................... | .................... | .................... | .................... |
| V | 12 | hCoV-19/Japan/PG-0801/2020\|EPI_ISL_684411\|2020-04 | .................... | .................... | .................... | .................... | .................... | .................... |
| V | 13 | hCoV-19/Japan/PG-0802/2020\|EPI_ISL_684412\|2020-04 | .................... | .................... | .................... | .................... | .................... | .................... |
| V | 14 | hCoV-19/Japan/PG-0803/2020\|EPI_ISL_684413\|2020-04 | .................... | .................... | .................... | .................... | .................... | .................... |
| V | 15 | hCoV-19/Japan/PG-0804/2020\|EPI_ISL_684414\|2020-04 | .................... | .................... | .................... | .................... | .................... | .................... |
| V | 16 | hCoV-19/Japan/PG-0805/2020\|EPI_ISL_684415\|2020-04 | .................... | .................... | .................... | .................... | .................... | .................... |
| V | 17 | hCoV-19/Japan/PG-0806/2020\|EPI_ISL_684416\|2020-04 | .................... | .................... | .................... | .................... | .................... | .................... |
| V | 18 | hCoV-19/Japan/PG-0807/2020\|EPI_ISL_684417\|2020-04 | .................... | .................... | .................... | .................... | .................... | .................... |
| V | 19 | hCoV-19/Japan/PG-0808/2020\|EPI_ISL_684418\|2020-04 | .................... | .................... | .................... | .................... | .................... | .................... |
| V | 20 | hCoV-19/Japan/PG-0809/2020\|EPI_ISL_684419\|2020-04 | .................... | .................... | .................... | .................... | .................... | .................... |
| V | 21 | hCoV-19/Japan/PG-0810/2020\|EPI_ISL_684420\|2020-04 | .................... | .................... | .................... | .................... | .................... | .................... |
| V | 22 | hCoV-19/Japan/PG-0811/2020\|EPI_ISL_684421\|2020-03 | .................... | .................... | .................... | .................... | .................... | .................... |
| V | 23 | hCoV-19/Japan/PG-0812/2020\|EPI_ISL_684422\|2020-03 | .................... | .................... | .................... | .................... | .................... | .................... |

| | | | | | | | |
|---|---|---|---|---|---|---|---|
| V | 24 hCoV-19/Japan/PG-0813/2020\|EPI_ISL_684423\|2020-04 | .................... | .................... | ..................... | .................... | .................... | .................... |
| V | 25 hCoV-19/Japan/PG-0814/2020\|EPI_ISL_684424\|2020-04 | .................... | .................... | ..................... | .................... | .................... | .................... |
| V | 26 hCoV-19/Japan/PG-0815/2020\|EPI_ISL_684425\|2020-04 | .................... | .................... | ..................... | .................... | .................... | .................... |
| V | 27 hCoV-19/Japan/PG-0816/2020\|EPI_ISL_684426\|2020-04 | .................... | .................... | ..................... | .................... | .................... | .................... |
| V | 28 hCoV-19/Japan/PG-0817/2020\|EPI_ISL_684427\|2020-04 | .................... | .................... | ..................... | .................... | .................... | .................... |
| V | 29 hCoV-19/Japan/PG-0818/2020\|EPI_ISL_684428\|2020-04 | .................... | .................... | ..................... | .................... | .................... | .................... |
| V | 30 hCoV-19/Japan/PG-0819/2020\|EPI_ISL_684429\|2020-03 | .................... | .................... | ..................... | .................... | .................... | .................... |
| V | 31 hCoV-19/Japan/PG-0820/2020\|EPI_ISL_684430\|2020-04 | .................... | .................... | ..................... | .................... | .................... | .................... |
| V | 32 hCoV-19/Japan/PG-0821/2020\|EPI_ISL_684431\|2020-04 | .................... | .................... | ..................... | .................... | .................... | .................... |
| V | 33 hCoV-19/Japan/PG-0822/2020\|EPI_ISL_684432\|2020-04 | .................... | .................... | ..................... | .................... | .................... | .................... |
| V | 34 hCoV-19/Japan/PG-0823/2020\|EPI_ISL_684433\|2020-04 | .................... | .................... | ..................... | .................... | .................... | .................... |
| V | 35 hCoV-19/Japan/PG-0824/2020\|EPI_ISL_684434\|2020-03 | .................... | .................... | ..................... | .................... | .................... | .................... |
| V | 36 hCoV-19/Japan/PG-0825/2020\|EPI_ISL_684435\|2020-03 | .................... | .................... | ..................... | .................... | .................... | .................... |
| V | 37 hCoV-19/Japan/PG-0826/2020\|EPI_ISL_684436\|2020-03 | .................... | .................... | ..................... | .................... | .................... | .................... |
| V | 38 hCoV-19/Japan/PG-0827/2020\|EPI_ISL_684437\|2020-03 | .................... | .................... | ..................... | .................... | .................... | .................... |
| V | 39 hCoV-19/Japan/PG-0828/2020\|EPI_ISL_684438\|2020-03 | .................... | .................... | ..................... | .................... | .................... | .................... |
| V | 40 hCoV-19/Japan/PG-0829/2020\|EPI_ISL_684439\|2020-03 | .................... | .................... | ..................... | .................... | .................... | .................... |
| V | 41 hCoV-19/Japan/PG-0830/2020\|EPI_ISL_684440\|2020-04 | .................... | .................... | ..................... | .................... | .................... | .................... |
| V | 42 hCoV-19/Japan/PG-0831/2020\|EPI_ISL_684441\|2020-04 | .................... | .................... | ..................... | .................... | .................... | .................... |
| V | 43 hCoV-19/Japan/PG-0832/2020\|EPI_ISL_684442\|2020-04 | .................... | .................... | ..................... | .................... | .................... | .................... |
| V | 44 hCoV-19/Japan/PG-0833/2020\|EPI_ISL_684443\|2020-04 | .................... | .................... | ..................... | .................... | .................... | .................... |
| V | 45 hCoV-19/Japan/PG-0834/2020\|EPI_ISL_684444\|2020-04 | .................... | .................... | ..................... | .................... | .................... | .................... |
| V | 46 hCoV-19/Japan/PG-0835/2020\|EPI_ISL_684445\|2020-04 | .................... | .................... | ..................... | .................... | .................... | .................... |
| V | 47 hCoV-19/Japan/PG-0836/2020\|EPI_ISL_684446\|2020-04 | .................... | .................... | ..................... | .................... | .................... | .................... |
| V | 48 hCoV-19/Japan/PG-0837/2020\|EPI_ISL_684447\|2020-04 | .................... | .................... | ..................... | .................... | .................... | .................... |
| V | 49 hCoV-19/Japan/PG-0838/2020\|EPI_ISL_684448\|2020-04 | .................... | .................... | ..................... | .................... | .................... | .................... |
| V | 50 hCoV-19/Japan/PG-0839/2020\|EPI_ISL_684449\|2020-04 | .................... | .................... | ..................... | .................... | .................... | .................... |
| V | 51 hCoV-19/Japan/PG-0840/2020\|EPI_ISL_684450\|2020-04 | .................... | .................... | ..................... | .................... | .................... | .................... |
| V | 52 hCoV-19/Japan/PG-0841/2020\|EPI_ISL_684451\|2020-04 | .................... | .................... | ..................... | .................... | .................... | .................... |
| V | 53 hCoV-19/Japan/PG-0842/2020\|EPI_ISL_684452\|2020-04 | .................... | .................... | ..................... | .................... | .................... | .................... |
| V | 54 hCoV-19/Japan/PG-0843/2020\|EPI_ISL_684453\|2020-04 | .................... | .................... | ..................... | .................... | .................... | .................... |
| V | 55 hCoV-19/Japan/PG-0844/2020\|EPI_ISL_684454\|2020-04 | .................... | .................... | ..................... | .................... | .................... | .................... |
| V | 56 hCoV-19/Japan/PG-0845/2020\|EPI_ISL_684455\|2020-04 | .................... | .................... | ..................... | .................... | .................... | .................... |
| V | 57 hCoV-19/Japan/PG-0846/2020\|EPI_ISL_684456\|2020-04 | .................... | .................... | ..................... | .................... | .................... | .................... |
| V | 58 hCoV-19/Japan/PG-0847/2020\|EPI_ISL_684457\|2020-04 | .................... | .................... | ..................... | .................... | .................... | .................... |
| V | 59 hCoV-19/Japan/PG-0848/2020\|EPI_ISL_684458\|2020-04 | .................... | .................... | ..................... | .................... | .................... | .................... |
| V | 60 hCoV-19/Japan/PG-0849/2020\|EPI_ISL_684459\|2020-04 | .................... | .................... | ..................... | .................... | .................... | .................... |
| V | 61 hCoV-19/Japan/PG-0850/2020\|EPI_ISL_684460\|2020-04 | .................... | .................... | ..................... | .................... | .................... | .................... |
| V | 62 hCoV-19/Japan/PG-0851/2020\|EPI_ISL_684461\|2020-04 | .................... | .................... | ..................... | .................... | .................... | .................... |
| V | 63 hCoV-19/Japan/PG-0852/2020\|EPI_ISL_684462\|2020-04 | .................... | .................... | ..................... | .................... | .................... | .................... |
| V | 64 hCoV-19/Japan/PG-0853/2020\|EPI_ISL_684463\|2020-04 | .................... | .................... | ..................... | .................... | .................... | .................... |
| V | 65 hCoV-19/Japan/PG-0854/2020\|EPI_ISL_684464\|2020-04 | .................... | .................... | ..................... | .................... | .................... | .................... |
| V | 66 hCoV-19/Japan/PG-0855/2020\|EPI_ISL_684465\|2020-04 | .................... | .................... | ..................... | .................... | .................... | .................... |
| V | 67 hCoV-19/Japan/PG-0856/2020\|EPI_ISL_684466\|2020-04 | .................... | .................... | ..................... | .................... | .................... | .................... |
| V | 68 hCoV-19/Japan/PG-0857/2020\|EPI_ISL_684467\|2020-04 | .................... | .................... | ..................... | .................... | .................... | .................... |
| V | 69 hCoV-19/Japan/PG-0858/2020\|EPI_ISL_684468\|2020-04 | .................... | .................... | ..................... | .................... | .................... | .................... |
| V | 70 hCoV-19/Japan/PG-0859/2020\|EPI_ISL_684469\|2020-04 | .................... | .................... | ..................... | .................... | .................... | .................... |
| V | 71 hCoV-19/Japan/PG-0860/2020\|EPI_ISL_684470\|2020-04 | .................... | .................... | ..................... | .................... | .................... | .................... |
| V | 72 hCoV-19/Japan/PG-0861/2020\|EPI_ISL_684471\|2020 | .................... | .................... | ..................... | .................... | .................... | .................... |
| V | 73 hCoV-19/Japan/PG-0862/2020\|EPI_ISL_684472\|2020 | .................... | .................... | ..................... | .................... | .................... | .................... |
| V | 74 hCoV-19/Japan/PG-0863/2020\|EPI_ISL_684473\|2020 | .................... | .................... | ..................... | .................... | .................... | .................... |
| V | 75 hCoV-19/Japan/PG-0864/2020\|EPI_ISL_684474\|2020 | .................... | .................... | ..................... | .................... | .................... | .................... |
| V | 76 hCoV-19/Japan/PG-0865/2020\|EPI_ISL_684475\|2020 | .................... | .................... | ..................... | .................... | .................... | .................... |
| V | 77 hCoV-19/Japan/PG-0866/2020\|EPI_ISL_684476\|2020 | .................... | .................... | ..................... | .................... | .................... | .................... |
| V | 78 hCoV-19/Japan/PG-0867/2020\|EPI_ISL_684477\|2020 | .................... | .................... | ..................... | .................... | .................... | .................... |
| V | 79 hCoV-19/Japan/PG-0868/2020\|EPI_ISL_684478\|2020 | .................... | .................... | ..................... | .................... | .................... | .................... |
| V | 80 hCoV-19/Japan/PG-0869/2020\|EPI_ISL_684479\|2020 | .................... | .................... | ..................... | .................... | .................... | .................... |
| V | 81 hCoV-19/Japan/PG-0870/2020\|EPI_ISL_684480\|2020 | .................... | .................... | ..................... | .................... | .................... | .................... |
| V | 82 hCoV-19/Japan/PG-0871/2020\|EPI_ISL_684481\|2020 | .................... | .................... | ..................... | .................... | .................... | .................... |
| V | 83 hCoV-19/Japan/PG-0872/2020\|EPI_ISL_684482\|2020 | .................... | .................... | ..................... | .................... | .................... | .................... |
| V | 84 hCoV-19/Japan/PG-0873/2020\|EPI_ISL_684483\|2020 | .................... | .................... | ..................... | .................... | .................... | .................... |
| V | 85 hCoV-19/Japan/PG-0874/2020\|EPI_ISL_684484\|2020-04 | .................... | .................... | ..................... | .................... | .................... | .................... |
| V | 86 hCoV-19/Japan/PG-0875/2020\|EPI_ISL_684485\|2020-04 | .................... | .................... | ..................... | .................... | .................... | .................... |
| V | 87 hCoV-19/Japan/PG-0876/2020\|EPI_ISL_684486\|2020-03 | .................... | .................... | ..................... | .................... | .................... | .................... |
| V | 88 hCoV-19/Japan/PG-0877/2020\|EPI_ISL_684487\|2020-03 | .................... | .................... | ..................... | .................... | .................... | .................... |
| V | 89 hCoV-19/Japan/PG-0878/2020\|EPI_ISL_684488\|2020-03 | .................... | .................... | ..................... | .................... | .................... | .................... |
| V | 90 hCoV-19/Japan/PG-0879/2020\|EPI_ISL_684489\|2020-04 | .................... | .................... | ..................... | .................... | .................... | .................... |
| V | 91 hCoV-19/Japan/PG-0880/2020\|EPI_ISL_684490\|2020-04 | .................... | .................... | ..................... | .................... | .................... | .................... |
| V | 92 hCoV-19/Japan/PG-0881/2020\|EPI_ISL_684491\|2020-04 | .................... | .................... | ..................... | .................... | .................... | .................... |
| V | 93 hCoV-19/Japan/PG-0882/2020\|EPI_ISL_684492\|2020-03 | .................... | .................... | ..................... | .................... | .................... | .................... |
| V | 94 hCoV-19/Japan/PG-0883/2020\|EPI_ISL_684493\|2020-03 | .................... | .................... | ..................... | .................... | .................... | .................... |
| V | 95 hCoV-19/Japan/PG-0884/2020\|EPI_ISL_684494\|2020-04 | .................... | .................... | ..................... | .................... | .................... | .................... |
| V | 96 hCoV-19/Japan/PG-0885/2020\|EPI_ISL_684495\|2020-04 | .................... | .................... | ..................... | .................... | .................... | .................... |
| V | 97 hCoV-19/Japan/PG-0886/2020\|EPI_ISL_684496\|2020-04 | .................... | .................... | ..................... | .................... | .................... | .................... |
| V | 98 hCoV-19/Japan/PG-0887/2020\|EPI_ISL_684497\|2020-04 | .................... | .................... | ..................... | .................... | .................... | .................... |
| V | 99 hCoV-19/Japan/PG-0888/2020\|EPI_ISL_684498\|2020-04 | .................... | .................... | ..................... | .................... | .................... | .................... |
| V | 100 hCoV-19/Japan/PG-0889/2020\|EPI_ISL_684499\|2020-04 | .................... | .................... | ..................... | .................... | .................... | .................... |
| V | 101 hCoV-19/Japan/PG-2095/2020\|EPI_ISL_685700\|2020-03 | .................... | .................... | ..................... | .................... | .................... | .................... |
| V | 102 hCoV-19/Japan/PG-2096/2020\|EPI_ISL_685701\|2020-04 | .................... | .................... | ..................... | .................... | .................... | .................... |
| V | 103 hCoV-19/Japan/PG-2097/2020\|EPI_ISL_685702\|2020-04 | .................... | .................... | ..................... | .................... | .................... | ...............A.... |

V 104 hCoV-19/Japan/PG-2098/2020|EPI_ISL_685703|2020-04 .................... .................... ..................... .................... .................... ....................
V 105 hCoV-19/Japan/PG-2099/2020|EPI_ISL_685704|2020-04 .................... .................... ..................... .................... .................... ....................
V 106 hCoV-19/Japan/PG-2100/2020|EPI_ISL_685705|2020-04 .................... .................... ..................... .................... .................... ....................
V 108 hCoV-19/Japan/PG-2102/2020|EPI_ISL_685707|2020-04 .................... .................... ..................... .................... .................... ....................
V 109 hCoV-19/Japan/PG-2103/2020|EPI_ISL_685708|2020-04 .................... .................... ..................... .................... .................... ....................
V 110 hCoV-19/Japan/PG-2104/2020|EPI_ISL_685709|2020-04 .................... .................... ..................... .................... .................... ....................
V 111 hCoV-19/Japan/PG-2105/2020|EPI_ISL_685710|2020-04 .................... .................... ..................... .................... .................... ....................
V 112 hCoV-19/Japan/PG-2106/2020|EPI_ISL_685711|2020-04 .................... .................... ..................... .................... .................... ....................
V 113 hCoV-19/Japan/PG-2107/2020|EPI_ISL_685712|2020-05 .................... .................... ..................... .................... .................... ....................
V 114 hCoV-19/Japan/PG-2108/2020|EPI_ISL_685713|2020-05 .................... .................... ..................... .................... .................... ....................
V 115 hCoV-19/Japan/PG-2109/2020|EPI_ISL_685714|2020-05 .................... .................... ..................... .................... .................... ....................
V 116 hCoV-19/Japan/PG-2110/2020|EPI_ISL_685715|2020-05 .................... .................... ..................... .................... .................... ....................
V 117 hCoV-19/Japan/PG-2111/2020|EPI_ISL_685716|2020-05 .................... .................... ..................... .................... .................... ....................
V 118 hCoV-19/Japan/PG-2112/2020|EPI_ISL_685717|2020-05 .................... .................... ..................... .................... .................... ....................
V 119 hCoV-19/Japan/PG-2113/2020|EPI_ISL_685718|2020-05 .................... .................... ..................... .................... .................... ....................
V 120 hCoV-19/Japan/PG-2114/2020|EPI_ISL_685719|2020-05 .................... .................... ..................... .................... .................... ....................
V 121 hCoV-19/Japan/PG-2115/2020|EPI_ISL_685720|2020-05 .................... .................... ..................... .................... .................... ....................
V 122 hCoV-19/Japan/PG-2116/2020|EPI_ISL_685721|2020-05 .................... ......A............. ..................... .................... .................... ....................
V 123 hCoV-19/Japan/PG-2117/2020|EPI_ISL_685722|2020-05 .................... .................... ..................... .................... .................... ....................
V 124 hCoV-19/Japan/PG-2118/2020|EPI_ISL_685723|2020-05 .................... ......A............. ..................... .................... .................... ....................
V 125 hCoV-19/Japan/PG-2119/2020|EPI_ISL_685724|2020-05 .................... .................... ..................... .................... .................... ....................
V 126 hCoV-19/Japan/PG-2120/2020|EPI_ISL_685725|2020-05 .................... .................... ..................... .................... .................... ....................
V 127 hCoV-19/Japan/PG-2121/2020|EPI_ISL_685726|2020-05 .................... .................... ..................... .................... .................... ....................
V 128 hCoV-19/Japan/PG-2122/2020|EPI_ISL_685727|2020-05 .................... .................... ..................... .................... .................... ....................
V 129 hCoV-19/Japan/PG-2123/2020|EPI_ISL_685728|2020-05 .................... .................... ..................... .................... .................... ....................
V 130 hCoV-19/Japan/PG-2124/2020|EPI_ISL_685729|2020-05 .................... ......A............. ..................... .................... .................... ....................
V 131 hCoV-19/Japan/PG-2125/2020|EPI_ISL_685730|2020-05 .................... .................... ..................... .................... .................... ....................
V 132 hCoV-19/Japan/PG-2126/2020|EPI_ISL_685731|2020-05 .................... .................... ..................... .................... .................... ....................
V 133 hCoV-19/Japan/PG-2127/2020|EPI_ISL_685732|2020-05 .................... .................... ..................... .................... .................... ....................
V 134 hCoV-19/Japan/PG-2128/2020|EPI_ISL_685733|2020-05 .................... .................... ..................... .................... .................... ....................
V 135 hCoV-19/Japan/PG-2129/2020|EPI_ISL_685734|2020-04 .................... .................... ..................... .................... .................... ....................
V 136 hCoV-19/Japan/PG-2130/2020|EPI_ISL_685735|2020-04 .................... .................... ..................... .................... .................... ....................
V 137 hCoV-19/Japan/PG-2131/2020|EPI_ISL_685736|2020-04 .................... .................... ..................... .................... .................... ....................
V 138 hCoV-19/Japan/PG-2132/2020|EPI_ISL_685737|2020-04 .................... .................... ..................... .................... .................... ....................
V 139 hCoV-19/Japan/PG-2133/2020|EPI_ISL_685738|2020-04 .................... .................... ..................... .................... .................... ....................
V 140 hCoV-19/Japan/PG-2134/2020|EPI_ISL_685739|2020-04 .................... .................... ..................... .................... .................... ....................
V 141 hCoV-19/Japan/PG-2135/2020|EPI_ISL_685740|2020-04 .................... .................... ..................... .................... .................... ....................
V 142 hCoV-19/Japan/PG-2136/2020|EPI_ISL_685741|2020-04 .................... .................... ..................... .................... .................... ....................
V 143 hCoV-19/Japan/PG-2137/2020|EPI_ISL_685742|2020-04 .................... .................... ..................... .................... .................... ....................
V 144 hCoV-19/Japan/PG-2138/2020|EPI_ISL_685743|2020-04 .................... .................... ..................... .................... .................... ....................
V 145 hCoV-19/Japan/PG-2139/2020|EPI_ISL_685744|2020-05 .................... .................... ..................... .................... .................... ....................
V 146 hCoV-19/Japan/PG-2140/2020|EPI_ISL_685745|2020-05 .................... .................... ..................... .................... .................... ....................
V 147 hCoV-19/Japan/PG-2141/2020|EPI_ISL_685746|2020-04 .................... .................... ..................... .................... .................... ....................
V 148 hCoV-19/Japan/PG-2142/2020|EPI_ISL_685747|2020-04 .................... .................... ..................... .................... .................... ....................
V 149 hCoV-19/Japan/PG-2143/2020|EPI_ISL_685748|2020-04 .................... .................... ..................... .................... .................... ....................
V 150 hCoV-19/Japan/PG-2144/2020|EPI_ISL_685749|2020-04 .................... .................... ..................... .................... .................... ....................
V 151 hCoV-19/Japan/PG-2145/2020|EPI_ISL_685750|2020-04 .................... .................... ..................... .................... .................... ....................
V 152 hCoV-19/Japan/PG-2146/2020|EPI_ISL_685751|2020-04 .................... .................... ..................... .................... .................... ....................
V 153 hCoV-19/Japan/PG-2147/2020|EPI_ISL_685752|2020-04 .................... .................... ..................... .................... .................... ....................
V 154 hCoV-19/Japan/PG-2148/2020|EPI_ISL_685753|2020-04 .................... .................... ..................... .................... .................... ....................
V 155 hCoV-19/Japan/PG-2149/2020|EPI_ISL_685754|2020-04 .................... .................... ..................... .................... .................... ....................
V 156 hCoV-19/Japan/PG-2150/2020|EPI_ISL_685755|2020-04 .................... .................... ..................... .................... .................... ....................
V 157 hCoV-19/Japan/PG-2151/2020|EPI_ISL_685756|2020-04 .................... .................... ..................... .................... .................... ....................
V 158 hCoV-19/Japan/PG-2152/2020|EPI_ISL_685757|2020-04 .................... .................... ..................... .................... .................... ....................
V 159 hCoV-19/Japan/PG-2153/2020|EPI_ISL_685758|2020-04 .................... .................... ..................... .................... .................... ....................
V 160 hCoV-19/Japan/PG-2154/2020|EPI_ISL_685759|2020-04 .................... .................... ..................... .................... .................... ....................
V 161 hCoV-19/Japan/PG-2155/2020|EPI_ISL_685760|2020-04 .................... .................... ..................... .................... .................... ....................
V 162 hCoV-19/Japan/PG-2156/2020|EPI_ISL_685761|2020-04 .................... .................... ..................... .................... .................... ....................
V 163 hCoV-19/Japan/PG-2157/2020|EPI_ISL_685762|2020-04 .................... .................... ..................... .................... .................... ....................
V 164 hCoV-19/Japan/PG-2158/2020|EPI_ISL_685763|2020-04 .................... .................... ..................... .................... .................... ....................
V 165 hCoV-19/Japan/PG-2159/2020|EPI_ISL_685764|2020-04 .................... .................... ..................... .................... .................... ....................
V 166 hCoV-19/Japan/PG-2160/2020|EPI_ISL_685765|2020-04 .................... .................... ..................... .................... .................... ....................
V 167 hCoV-19/Japan/PG-2161/2020|EPI_ISL_685766|2020-04 .................... .................... ..................... .................... .................... ....................
V 168 hCoV-19/Japan/PG-2162/2020|EPI_ISL_685767|2020-04 .................... .................... ..................... .................... .................... ....................
V 169 hCoV-19/Japan/PG-2163/2020|EPI_ISL_685768|2020-04 .................... .................... ..................... .................... .................... ....................
V 170 hCoV-19/Japan/PG-2164/2020|EPI_ISL_685769|2020-04 .................... .................... ..................... .................... .................... ....................
V 171 hCoV-19/Japan/PG-2165/2020|EPI_ISL_685770|2020-04 .................... .................... ..................... .................... .................... ....................
V 172 hCoV-19/Japan/PG-2166/2020|EPI_ISL_685771|2020-04 .................... .................... ..................... .................... .................... ....................
V 173 hCoV-19/Japan/PG-2167/2020|EPI_ISL_685772|2020-04 .................... .................... ..................... .................... .................... ....................
V 174 hCoV-19/Japan/PG-2168/2020|EPI_ISL_685773|2020-04 .................... .................... ..................... .................... .................... ....................
V 175 hCoV-19/Japan/PG-2169/2020|EPI_ISL_685774|2020-04 .................... .................... ..................... .................... .................... ....................
V 176 hCoV-19/Japan/PG-2170/2020|EPI_ISL_685775|2020-05 .................... .................... ..................... .................... .................... ....................
V 177 hCoV-19/Japan/PG-2171/2020|EPI_ISL_685776|2020-05 .................... .................... ..................... .................... .................... ....................
V 178 hCoV-19/Japan/PG-2172/2020|EPI_ISL_685777|2020-05 .................... .................... ..................... .................... .................... ....................
V 179 hCoV-19/Japan/PG-2173/2020|EPI_ISL_685778|2020-05 .................... .................... ..................... .................... .................... ....................
V 180 hCoV-19/Japan/PG-2174/2020|EPI_ISL_685779|2020-05 .................... .................... ..................... .................... .................... ....................
V 181 hCoV-19/Japan/PG-2175/2020|EPI_ISL_685780|2020-02 .................... .................... ..................... .................... .................... ....................
V 182 hCoV-19/Japan/PG-2176/2020|EPI_ISL_685781|2020-03 .................... .................... ..................... .................... .................... ....................
V 183 hCoV-19/Japan/PG-2177/2020|EPI_ISL_685782|2020-03 .................... .................... ..................... .................... .................... ....................
V 184 hCoV-19/Japan/PG-2178/2020|EPI_ISL_685783|2020-03 .................... .................... ..................... .................... .................... ....................

| | | | | | | | |
|---|---|---|---|---|---|---|---|
| V | 185 hCoV-19/Japan/PG-2179/2020\|EPI_ISL_685784\|2020-03 | ···················· | ···················· | ···················· | ···················· | ···················· | ···················· |
| V | 186 hCoV-19/Japan/PG-2180/2020\|EPI_ISL_685785\|2020-03 | ···················· | ···················· | ···················· | ···················· | ···················· | ···················· |
| V | 187 hCoV-19/Japan/PG-2181/2020\|EPI_ISL_685786\|2020-04 | ···················· | ···················· | ···················· | ···················· | ···················· | ···················· |
| V | 188 hCoV-19/Japan/PG-2182/2020\|EPI_ISL_685787\|2020-04 | ···················· | ···················· | ···················· | ···················· | ···················· | ···················· |
| V | 189 hCoV-19/Japan/PG-2183/2020\|EPI_ISL_685788\|2020-04 | ···················· | ···················· | ···················· | ···················· | ···················· | ···················· |
| V | 190 hCoV-19/Japan/PG-2184/2020\|EPI_ISL_685789\|2020-04 | ···················· | ···················· | ···················· | ···················· | ···················· | ···················· |
| V | 191 hCoV-19/Japan/PG-2185/2020\|EPI_ISL_685790\|2020-04 | ···················· | ···················· | ···················· | ···················· | ···················· | ···················· |
| V | 192 hCoV-19/Japan/PG-2186/2020\|EPI_ISL_685791\|2020-04 | ···················· | ···················· | ···················· | ···················· | ···················· | ···················· |
| V | 193 hCoV-19/Japan/PG-2187/2020\|EPI_ISL_685792\|2020-04 | ···················· | ···················· | ···················· | ···················· | ···················· | ···················· |
| V | 194 hCoV-19/Japan/PG-2188/2020\|EPI_ISL_685793\|2020-04 | ···················· | ···················· | ···················· | ···················· | ···················· | ···················· |
| V | 195 hCoV-19/Japan/PG-2189/2020\|EPI_ISL_685794\|2020-04 | ···················· | ···················· | ···················· | ···················· | ···················· | ···················· |
| V | 196 hCoV-19/Japan/PG-2190/2020\|EPI_ISL_685795\|2020-04 | ···················· | ···················· | ···················· | ···················· | ···················· | ···················· |
| V | 197 hCoV-19/Japan/PG-2191/2020\|EPI_ISL_685796\|2020-04 | ···················· | ···················· | ···················· | ···················· | ···················· | ···················· |
| V | 198 hCoV-19/Japan/PG-2192/2020\|EPI_ISL_685797\|2020-04 | ···················· | ···················· | ···················· | ···················· | ···················· | ···················· |
| V | 199 hCoV-19/Japan/PG-2193/2020\|EPI_ISL_685798\|2020-04 | ···················· | ···················· | ···················· | ···················· | ···················· | ···················· |
| V | 200 hCoV-19/Japan/PG-2194/2020\|EPI_ISL_685799\|2020-04 | ···················· | ···················· | ···················· | ···················· | ···················· | ···················· |
| V | 201 hCoV-19/Japan/PG-6088/2020\|EPI_ISL_689200\|2020-07 | ···················· | ···················· | ···················· | ···················· | ···················· | ···················· |
| V | 202 hCoV-19/Japan/PG-6089/2020\|EPI_ISL_689201\|2020-07 | ···················· | ···················· | ···················· | ···················· | ···················· | ···················· |
| V | 203 hCoV-19/Japan/PG-6090/2020\|EPI_ISL_689202\|2020-07 | ···················· | ···················· | ···················· | ···················· | ···················· | ···················· |
| V | 204 hCoV-19/Japan/PG-6091/2020\|EPI_ISL_689203\|2020-08 | ···················· | ···················· | ···················· | ···················· | ···················· | ···················· |
| V | 205 hCoV-19/Japan/PG-6092/2020\|EPI_ISL_689204\|2020-08 | ···················· | ···················· | ···················· | ···················· | ···················· | ···················· |
| V | 206 hCoV-19/Japan/PG-6093/2020\|EPI_ISL_689205\|2020-08 | ···················· | ···················· | ···················· | ···················· | ···················· | ···················· |
| V | 207 hCoV-19/Japan/PG-6094/2020\|EPI_ISL_689206\|2020-08 | ···················· | ···················· | ···················· | ···················· | ···················· | ···················· |
| V | 208 hCoV-19/Japan/PG-6178/2020\|EPI_ISL_689290\|2020-08 | ···················· | ···················· | ···················· | ···················· | ···················· | ···················· |
| V | 209 hCoV-19/Japan/PG-6179/2020\|EPI_ISL_689291\|2020-08 | ···················· | ···················· | ···················· | ···················· | ···················· | ···················· |
| V | 210 hCoV-19/Japan/PG-6180/2020\|EPI_ISL_689292\|2020-08 | ···················· | ···················· | ···················· | ···················· | ···················· | ···················· |
| V | 211 hCoV-19/Japan/PG-6181/2020\|EPI_ISL_689293\|2020-08 | ···················· | ···················· | ···················· | ···················· | ···················· | ···················· |
| V | 212 hCoV-19/Japan/PG-6182/2020\|EPI_ISL_689294\|2020-08 | ···················· | ···················· | ···················· | ···················· | ···················· | ···················· |
| V | 213 hCoV-19/Japan/PG-6183/2020\|EPI_ISL_689295\|2020-08 | ···················· | ···················· | ···················· | ···················· | ···················· | ···················· |
| V | 214 hCoV-19/Japan/PG-6184/2020\|EPI_ISL_689296\|2020-08 | ···················· | ···················· | ···················· | ···················· | ···················· | ···················· |
| V | 215 hCoV-19/Japan/PG-6185/2020\|EPI_ISL_689297\|2020-08 | ···················· | ···················· | ···················· | ···················· | ···················· | ···················· |
| V | 216 hCoV-19/Japan/PG-6186/2020\|EPI_ISL_689298\|2020-08 | ···················· | ···················· | ···················· | ···················· | ···················· | ···················· |
| V | 217 hCoV-19/Japan/PG-6187/2020\|EPI_ISL_689299\|2020-08 | ···················· | ···················· | ···················· | ···················· | ···················· | ···················· |
| V | 218 hCoV-19/Japan/PG-5171/2020\|EPI_ISL_690274\|2020-08 | ···················· | ···················· | ···················· | ···················· | ···················· | ···················· |
| V | 219 hCoV-19/Japan/PG-5172/2020\|EPI_ISL_690275\|2020-08 | ···················· | ···················· | ···················· | ···················· | ···················· | ···················· |
| V | 220 hCoV-19/Japan/PG-5173/2020\|EPI_ISL_690276\|2020-08 | ···················· | ···················· | ···················· | ···················· | ···················· | ···················· |
| V | 221 hCoV-19/Japan/PG-5174/2020\|EPI_ISL_690277\|2020-08 | ···················· | ···················· | ···················· | ···················· | ···················· | ···················· |
| V | 222 hCoV-19/Japan/PG-5175/2020\|EPI_ISL_690278\|2020-08 | ···················· | ···················· | ···················· | ···················· | ···················· | ···················· |
| V | 223 hCoV-19/Japan/PG-5176/2020\|EPI_ISL_690279\|2020-08 | ···················· | ···················· | ···················· | ···················· | ···················· | ···················· |
| V | 224 hCoV-19/Japan/PG-5177/2020\|EPI_ISL_690280\|2020-08 | ···················· | ···················· | ···················· | ···················· | ···················· | ···················· |
| V | 225 hCoV-19/Japan/PG-5178/2020\|EPI_ISL_690281\|2020-08 | ···················· | ···················· | ···················· | ···················· | ···················· | ···················· |
| V | 226 hCoV-19/Japan/PG-5179/2020\|EPI_ISL_690282\|2020-08 | ···················· | ···················· | ···················· | ···················· | ···················· | ···················· |
| V | 227 hCoV-19/Japan/PG-5180/2020\|EPI_ISL_690283\|2020-08 | ···················· | ···················· | ···················· | ···················· | ···················· | ···················· |
| V | 228 hCoV-19/Japan/PG-5181/2020\|EPI_ISL_690284\|2020-08 | ···················· | ···················· | ···················· | ···················· | ···················· | ···················· |
| V | 229 hCoV-19/Japan/PG-5182/2020\|EPI_ISL_690285\|2020-08 | ···················· | ···················· | ···················· | ···················· | ···················· | ···················· |
| V | 230 hCoV-19/Japan/PG-5183/2020\|EPI_ISL_690286\|2020-08 | ···················· | ···················· | ···················· | ···················· | ···················· | ···················· |
| V | 231 hCoV-19/Japan/PG-5184/2020\|EPI_ISL_690287\|2020-08 | ···················· | ···················· | ···················· | ···················· | ···················· | ···················· |
| V | 232 hCoV-19/Japan/PG-5185/2020\|EPI_ISL_690288\|2020-08 | ···················· | ···················· | ···················· | ···················· | ···················· | ···················· |
| V | 233 hCoV-19/Japan/PG-5186/2020\|EPI_ISL_690289\|2020-08 | ···················· | ···················· | ···················· | ···················· | ···················· | ···················· |
| V | 234 hCoV-19/Japan/PG-5187/2020\|EPI_ISL_690290\|2020-08 | ···················· | ···················· | ···················· | ···················· | ···················· | ···················· |
| V | 235 hCoV-19/Japan/PG-5188/2020\|EPI_ISL_690291\|2020-08 | ···················· | ···················· | ···················· | ···················· | ···················· | ···················· |
| V | 236 hCoV-19/Japan/PG-5189/2020\|EPI_ISL_690292\|2020-08 | ···················· | ···················· | ···················· | ···················· | ···················· | ···················· |
| V | 237 hCoV-19/Japan/PG-5190/2020\|EPI_ISL_690293\|2020-08 | ···················· | ···················· | ···················· | ···················· | ···················· | ···················· |
| V | 238 hCoV-19/Japan/PG-5191/2020\|EPI_ISL_690294\|2020-08 | ···················· | ···················· | ···················· | ···················· | ···················· | ···················· |
| V | 239 hCoV-19/Japan/PG-5192/2020\|EPI_ISL_690295\|2020-08 | ···················· | ···················· | ···················· | ···················· | ···················· | ···················· |
| V | 240 hCoV-19/Japan/PG-5193/2020\|EPI_ISL_690296\|2020-08 | ···················· | ···················· | ···················· | ···················· | ···················· | ···················· |
| V | 241 hCoV-19/Japan/PG-5194/2020\|EPI_ISL_690297\|2020-08 | ···················· | ···················· | ···················· | ···················· | ···················· | ···················· |
| V | 243 hCoV-19/Japan/PG-5196/2020\|EPI_ISL_690299\|2020-08 | ···················· | ···················· | ···················· | ···················· | ···················· | ···················· |
| V | 244 hCoV-19/Japan/PG-3406/2020\|EPI_ISL_691520\|2020-04 | ···················· | ···················· | ···················· | ···················· | ···················· | ···················· |
| V | 245 hCoV-19/Japan/PG-3407/2020\|EPI_ISL_691521\|2020-04 | ···················· | ···················· | ···················· | ···················· | ···················· | ···················· |
| V | 246 hCoV-19/Japan/PG-3409/2020\|EPI_ISL_691523\|2020-04 | ···················· | ···················· | ···················· | ···················· | ···················· | ···················· |
| V | 247 hCoV-19/Japan/PG-3410/2020\|EPI_ISL_691524\|2020-04 | ···················· | ···················· | ···················· | ···················· | ···················· | ···················· |
| V | 248 hCoV-19/Japan/PG-3411/2020\|EPI_ISL_691525\|2020-04 | ···················· | ···················· | ···················· | ···················· | ···················· | ···················· |
| V | 249 hCoV-19/Japan/PG-3412/2020\|EPI_ISL_691526\|2020-04 | ···················· | ···················· | ···················· | ···················· | ···················· | ···················· |
| V | 250 hCoV-19/Japan/PG-3413/2020\|EPI_ISL_691527\|2020-04 | ···················· | ···················· | ···················· | ···················· | ···················· | ···················· |
| V | 251 hCoV-19/Japan/PG-3414/2020\|EPI_ISL_691528\|2020-04 | ···················· | ···················· | ···················· | ···················· | ···················· | ···················· |
| V | 252 hCoV-19/Japan/PG-3417/2020\|EPI_ISL_691529\|2020-04 | ···················· | ···················· | ···················· | ···················· | ···················· | ···················· |
| V | 253 hCoV-19/USA/MA-MASPHL-00882/2020\|EPI_ISL_692850\|20 | ···················· | ···················· | ···················· | ···················· | ···················· | ···················· |
| V | 254 hCoV-19/USA/MA-MASPHL-00881/2020\|EPI_ISL_692851\|20 | ···················· | ·A·················· | ···················· | ···················· | ···················· | ···················· |
| V | 255 hCoV-19/USA/MA-MASPHL-00877/2020\|EPI_ISL_692855\|20 | ···················· | ···················· | ···················· | ···················· | ···················· | ···················· |
| V | 256 hCoV-19/USA/MA-MASPHL-00867/2020\|EPI_ISL_692858\|20 | ···················· | ···················· | ···················· | ···················· | ···················· | ···················· |
| V | 257 hCoV-19/Japan/PG-6111/2020\|EPI_ISL_689223\|2020-08 | ···················· | ···················· | ···················· | ···················· | ···················· | ···················· |
| V | 258 hCoV-19/Japan/PG-6112/2020\|EPI_ISL_689224\|2020-08 | ···················· | ···················· | ···················· | ···················· | ···················· | ···················· |
| V | 259 hCoV-19/Japan/PG-6113/2020\|EPI_ISL_689225\|2020-08 | ···················· | ···················· | ···················· | ···················· | ···················· | ···················· |
| V | 260 hCoV-19/Japan/PG-6114/2020\|EPI_ISL_689226\|2020-08 | ···················· | ···················· | ···················· | ···················· | ···················· | ···················· |
| V | 261 hCoV-19/Japan/PG-6115/2020\|EPI_ISL_689227\|2020-08 | ···················· | ···················· | ···················· | ···················· | ···················· | ···················· |
| V | 262 hCoV-19/Japan/PG-6116/2020\|EPI_ISL_689228\|2020-08 | ···················· | ···················· | ···················· | ···················· | ···················· | ···················· |
| V | 263 hCoV-19/Japan/PG-6117/2020\|EPI_ISL_689229\|2020-08 | ···················· | ···················· | ···················· | ···················· | ···················· | ···················· |
| V | 264 hCoV-19/Japan/PG-6118/2020\|EPI_ISL_689230\|2020-08 | ···················· | ···················· | ···················· | ···················· | ···················· | ···················· |
| V | 265 hCoV-19/Japan/PG-6119/2020\|EPI_ISL_689231\|2020-08 | ···················· | ···················· | ···················· | ···················· | ···················· | ···················· |

| | | | | | | | |
|---|---|---|---|---|---|---|---|
| V | 266 hCoV-19/Japan/PG-6120/2020\|EPI_ISL_689232\|2020-08 | ···················· | ···················· | ····················· | ···················· | ···················· | ···················· |
| V | 267 hCoV-19/Japan/PG-6121/2020\|EPI_ISL_689233\|2020-08 | ···················· | ···················· | ····················· | ···················· | ···················· | ···················· |
| V | 268 hCoV-19/Japan/PG-6122/2020\|EPI_ISL_689234\|2020-08 | ···················· | ····A··············· | ····················· | ···················· | ···················· | ···················· |
| V | 269 hCoV-19/Japan/PG-6123/2020\|EPI_ISL_689235\|2020-08 | ···················· | ···················· | ····················· | ···················· | ···················· | ···················· |
| V | 270 hCoV-19/Japan/PG-6124/2020\|EPI_ISL_689236\|2020-08 | ···················· | ···················· | ····················· | ···················· | ···················· | ···················· |
| V | 271 hCoV-19/Japan/PG-6125/2020\|EPI_ISL_689237\|2020-08 | ···················· | ···················· | ····················· | ···················· | ···················· | ···················· |
| V | 272 hCoV-19/Japan/PG-6126/2020\|EPI_ISL_689238\|2020-08 | ···················· | ···················· | ····················· | ···················· | ···················· | ···················· |
| V | 273 hCoV-19/Japan/PG-6127/2020\|EPI_ISL_689239\|2020-08 | ···················· | ···················· | ····················· | ···················· | ···················· | ···················· |
| V | 274 hCoV-19/Japan/PG-6128/2020\|EPI_ISL_689240\|2020-08 | ···················· | ···················· | ····················· | ···················· | ···················· | ···················· |
| V | 275 hCoV-19/Japan/PG-6129/2020\|EPI_ISL_689241\|2020-08 | ···················· | ···················· | ····················· | ···················· | ···················· | ···················· |
| V | 276 hCoV-19/Japan/PG-6130/2020\|EPI_ISL_689242\|2020-08 | ···················· | ···················· | ····················· | ···················· | ···················· | ···················· |
| V | 277 hCoV-19/Japan/PG-6131/2020\|EPI_ISL_689243\|2020-08 | ···················· | ···················· | ····················· | ···················· | ···················· | ···················· |
| V | 278 hCoV-19/Japan/PG-6132/2020\|EPI_ISL_689244\|2020-08 | ···················· | ···················· | ····················· | ···················· | ···················· | ···················· |
| V | 279 hCoV-19/Japan/PG-6133/2020\|EPI_ISL_689245\|2020-08 | ···················· | ···················· | ····················· | ···················· | ···················· | ···················· |
| V | 280 hCoV-19/Japan/PG-6134/2020\|EPI_ISL_689246\|2020-08 | ···················· | ···················· | ····················· | ···················· | ···················· | ···················· |
| V | 281 hCoV-19/Japan/PG-6135/2020\|EPI_ISL_689247\|2020-08 | ···················· | ···················· | ····················· | ···················· | ···················· | ···················· |
| V | 282 hCoV-19/Japan/PG-6136/2020\|EPI_ISL_689248\|2020-08 | ···················· | ···················· | ····················· | ···················· | ···················· | ···················· |
| V | 283 hCoV-19/Japan/PG-6137/2020\|EPI_ISL_689249\|2020-08 | ···················· | ···················· | ····················· | ···················· | ···················· | ···················· |
| V | 284 hCoV-19/Japan/PG-6138/2020\|EPI_ISL_689250\|2020-08 | ···················· | ···················· | ····················· | ···················· | ···················· | ···················· |
| V | 285 hCoV-19/Japan/PG-6139/2020\|EPI_ISL_689251\|2020-08 | ···················· | ···················· | ····················· | ···················· | ···················· | ···················· |
| V | 286 hCoV-19/Japan/PG-6140/2020\|EPI_ISL_689252\|2020-08 | ···················· | ···················· | ····················· | ···················· | ···················· | ···················· |
| V | 287 hCoV-19/Japan/PG-6141/2020\|EPI_ISL_689253\|2020-08 | ···················· | ···················· | ····················· | ···················· | ···················· | ···················· |
| V | 288 hCoV-19/Japan/PG-6142/2020\|EPI_ISL_689254\|2020-08 | ···················· | ···················· | ····················· | ···················· | ···················· | ···················· |
| V | 289 hCoV-19/Japan/PG-6143/2020\|EPI_ISL_689255\|2020-08 | ···················· | ···················· | ····················· | ···················· | ···················· | ···················· |
| V | 290 hCoV-19/Japan/PG-6144/2020\|EPI_ISL_689256\|2020-08 | ···················· | ···················· | ····················· | ···················· | ···················· | ···················· |
| V | 291 hCoV-19/Japan/PG-6145/2020\|EPI_ISL_689257\|2020-08 | ···················· | ···················· | ····················· | ···················· | ···················· | ···················· |
| V | 292 hCoV-19/Japan/PG-6146/2020\|EPI_ISL_689258\|2020-08 | ···················· | ···················· | ····················· | ···················· | ···················· | ···················· |
| V | 293 hCoV-19/Japan/PG-6147/2020\|EPI_ISL_689259\|2020-08 | ···················· | ···················· | ····················· | ···················· | ···················· | ···················· |
| V | 294 hCoV-19/Japan/PG-6148/2020\|EPI_ISL_689260\|2020-08 | ···················· | ···················· | ····················· | ···················· | ···················· | ···················· |
| V | 295 hCoV-19/Japan/PG-6149/2020\|EPI_ISL_689261\|2020-08 | ···················· | ···················· | ····················· | ···················· | ···················· | ···················· |
| V | 296 hCoV-19/Japan/PG-6150/2020\|EPI_ISL_689262\|2020-08 | ···················· | ···················· | ····················· | ···················· | ···················· | ···················· |
| V | 297 hCoV-19/Japan/PG-6151/2020\|EPI_ISL_689263\|2020-08 | ···················· | ···················· | ····················· | ···················· | ···················· | ···················· |
| V | 298 hCoV-19/Japan/PG-6152/2020\|EPI_ISL_689264\|2020-08 | ···················· | ···················· | ····················· | ···················· | ···················· | ···················· |
| V | 299 hCoV-19/Japan/PG-6153/2020\|EPI_ISL_689265\|2020-08 | ···················· | ···················· | ····················· | ···················· | ···················· | ···················· |
| V | 300 hCoV-19/Japan/PG-6154/2020\|EPI_ISL_689266\|2020-08 | ···················· | ···················· | ····················· | ···················· | ···················· | ···················· |
| V | 301 hCoV-19/Japan/PG-6155/2020\|EPI_ISL_689267\|2020-08 | ···················· | ···················· | ····················· | ···················· | ···················· | ···················· |
| V | 302 hCoV-19/Japan/PG-6156/2020\|EPI_ISL_689268\|2020-08 | ···················· | ···················· | ····················· | ···················· | ···················· | ···················· |
| V | 303 hCoV-19/Japan/PG-6157/2020\|EPI_ISL_689269\|2020-08 | ···················· | ···················· | ····················· | ···················· | ···················· | ···················· |
| V | 304 hCoV-19/Japan/PG-6158/2020\|EPI_ISL_689270\|2020-08 | ···················· | ···················· | ····················· | ···················· | ···················· | ···················· |
| V | 305 hCoV-19/Japan/PG-6159/2020\|EPI_ISL_689271\|2020-08 | ···················· | ···················· | ····················· | ···················· | ···················· | ···················· |
| V | 306 hCoV-19/Japan/PG-6160/2020\|EPI_ISL_689272\|2020-08 | ···················· | ···················· | ····················· | ···················· | ···················· | ···················· |
| V | 307 hCoV-19/Japan/PG-6161/2020\|EPI_ISL_689273\|2020-08 | ···················· | ···················· | ····················· | ···················· | ···················· | ···················· |
| V | 308 hCoV-19/Japan/PG-6162/2020\|EPI_ISL_689274\|2020-08 | ···················· | ···················· | ····················· | ···················· | ···················· | ···················· |
| V | 309 hCoV-19/Japan/PG-6163/2020\|EPI_ISL_689275\|2020-08 | ···················· | ···················· | ····················· | ···················· | ···················· | ···················· |
| V | 310 hCoV-19/Japan/PG-6164/2020\|EPI_ISL_689276\|2020-08 | ···················· | ···················· | ····················· | ···················· | ···················· | ···················· |
| V | 311 hCoV-19/Japan/PG-6165/2020\|EPI_ISL_689277\|2020-08 | ···················· | ···················· | ····················· | ···················· | ···················· | ···················· |
| V | 312 hCoV-19/Japan/PG-6166/2020\|EPI_ISL_689278\|2020-08 | ···················· | ···················· | ····················· | ···················· | ···················· | ···················· |
| V | 313 hCoV-19/Japan/PG-6167/2020\|EPI_ISL_689279\|2020-08 | ···················· | ···················· | ····················· | ···················· | ···················· | ···················· |
| V | 314 hCoV-19/Japan/PG-6168/2020\|EPI_ISL_689280\|2020-08 | ···················· | ···················· | ····················· | ···················· | ···················· | ···················· |
| V | 315 hCoV-19/Japan/PG-6169/2020\|EPI_ISL_689281\|2020-08 | ···················· | ···················· | ····················· | ···················· | ···················· | ···················· |
| V | 316 hCoV-19/Japan/PG-6170/2020\|EPI_ISL_689282\|2020-08 | ···················· | ···················· | ····················· | ···················· | ···················· | ···················· |
| V | 317 hCoV-19/Japan/PG-6171/2020\|EPI_ISL_689283\|2020-08 | ···················· | ···················· | ····················· | ···················· | ···················· | ···················· |
| V | 318 hCoV-19/Japan/PG-6172/2020\|EPI_ISL_689284\|2020-08 | ···················· | ···················· | ····················· | ···················· | ···················· | ···················· |
| V | 319 hCoV-19/Japan/PG-6173/2020\|EPI_ISL_689285\|2020-08 | ···················· | ···················· | ····················· | ···················· | ···················· | ···················· |
| V | 320 hCoV-19/Japan/PG-6174/2020\|EPI_ISL_689286\|2020-08 | ···················· | ···················· | ····················· | ···················· | ···················· | ···················· |
| V | 321 hCoV-19/Japan/PG-6175/2020\|EPI_ISL_689287\|2020-08 | ···················· | ···················· | ····················· | ···················· | ···················· | ···················· |
| V | 322 hCoV-19/Japan/PG-6176/2020\|EPI_ISL_689288\|2020-08 | ···················· | ···················· | ····················· | ···················· | ···················· | ···················· |
| V | 323 hCoV-19/Japan/PG-6177/2020\|EPI_ISL_689289\|2020-08 | ···················· | ···················· | ····················· | ···················· | ···················· | ···················· |
| V | 324 hCoV-19/Japan/PG-5090/2020\|EPI_ISL_690200\|2020-07 | ···················· | ···················· | ····················· | ···················· | ···················· | ···················· |
| V | 325 hCoV-19/Japan/PG-5091/2020\|EPI_ISL_690201\|2020-07 | ···················· | ···················· | ····················· | ···················· | ···················· | ···················· |
| V | 326 hCoV-19/Japan/PG-5092/2020\|EPI_ISL_690202\|2020-07 | ···················· | ···················· | ····················· | ···················· | ···················· | ···················· |
| V | 327 hCoV-19/Japan/PG-5093/2020\|EPI_ISL_690203\|2020-07 | ···················· | ···················· | ····················· | ···················· | ···················· | ···················· |
| V | 328 hCoV-19/Japan/PG-5094/2020\|EPI_ISL_690204\|2020-07 | ···················· | ···················· | ····················· | ···················· | ···················· | ···················· |
| V | 329 hCoV-19/Japan/PG-5095/2020\|EPI_ISL_690205\|2020-07 | ···················· | ···················· | ····················· | ···················· | ···················· | ···················· |
| V | 330 hCoV-19/Japan/PG-5096/2020\|EPI_ISL_690206\|2020-08 | ···················· | ···················· | ····················· | ···················· | ···················· | ···················· |
| V | 331 hCoV-19/Japan/PG-5097/2020\|EPI_ISL_690207\|2020-08 | ···················· | ···················· | ····················· | ···················· | ···················· | ···················· |
| V | 332 hCoV-19/Japan/PG-5098/2020\|EPI_ISL_690208\|2020-08 | ···················· | ···················· | ····················· | ···················· | ···················· | ···················· |
| V | 333 hCoV-19/Japan/PG-5099/2020\|EPI_ISL_690209\|2020-08 | ···················· | ···················· | ····················· | ···················· | ···················· | ···················· |
| V | 334 hCoV-19/Japan/PG-5101/2020\|EPI_ISL_690210\|2020-08 | ···················· | ···················· | ····················· | ···················· | ···················· | ···················· |
| V | 335 hCoV-19/Japan/PG-5102/2020\|EPI_ISL_690211\|2020-08 | ···················· | ···················· | ····················· | ···················· | ···················· | ···················· |
| V | 336 hCoV-19/Japan/PG-5103/2020\|EPI_ISL_690212\|2020-08 | ···················· | ···················· | ····················· | ···················· | ···················· | ···················· |
| V | 337 hCoV-19/Japan/PG-5104/2020\|EPI_ISL_690213\|2020-08 | ···················· | ···················· | ····················· | ···················· | ···················· | ···················· |
| V | 338 hCoV-19/Japan/PG-5105/2020\|EPI_ISL_690214\|2020-08 | ···················· | ···················· | ····················· | ···················· | ···················· | ···················· |
| V | 339 hCoV-19/Japan/PG-5106/2020\|EPI_ISL_690215\|2020-08 | ···················· | ···················· | ····················· | ···················· | ···················· | ···················· |
| V | 340 hCoV-19/Japan/PG-5107/2020\|EPI_ISL_690216\|2020-08 | ···················· | ···················· | ····················· | ···················· | ···················· | ···················· |
| V | 341 hCoV-19/Japan/PG-5108/2020\|EPI_ISL_690217\|2020-08 | ···················· | ···················· | ····················· | ···················· | ···················· | ···················· |
| V | 342 hCoV-19/Japan/PG-5109/2020\|EPI_ISL_690218\|2020-08 | ···················· | ···················· | ····················· | ···················· | ···················· | ···················· |
| V | 343 hCoV-19/Japan/PG-5110/2020\|EPI_ISL_690219\|2020-08 | ···················· | ···················· | ····················· | ···················· | ···················· | ···················· |
| V | 344 hCoV-19/Japan/PG-5111/2020\|EPI_ISL_690220\|2020-08 | ···················· | ···················· | ····················· | ···················· | ···················· | ···················· |
| V | 345 hCoV-19/Japan/PG-5112/2020\|EPI_ISL_690221\|2020-08 | ···················· | ···················· | ····················· | ···················· | ···················· | ···················· |

| | | | | | | | | |
|---|---|---|---|---|---|---|---|---|
| V | 346 | hCoV-19/Japan/PG-5113/2020\|EPI_ISL_690222\|2020-08 | ···················· | ···················· | ····················· | ···················· | ···················· | ···················· |
| V | 347 | hCoV-19/Japan/PG-5114/2020\|EPI_ISL_690223\|2020-08 | ···················· | ···················· | ····················· | ···················· | ···················· | ···················· |
| V | 348 | hCoV-19/Japan/PG-5115/2020\|EPI_ISL_690224\|2020-08 | ···················· | ···················· | ····················· | ···················· | ···················· | ···················· |
| V | 349 | hCoV-19/Japan/PG-5116/2020\|EPI_ISL_690225\|2020-08 | ···················· | ···················· | ····················· | ···················· | ···················· | ···················· |
| V | 350 | hCoV-19/Japan/PG-5117/2020\|EPI_ISL_690226\|2020-08 | ···················· | ···················· | ····················· | ···················· | ···················· | ···················· |
| V | 351 | hCoV-19/Japan/PG-5118/2020\|EPI_ISL_690227\|2020-08 | ···················· | ···················· | ····················· | ···················· | ···················· | ···················· |
| V | 352 | hCoV-19/Japan/PG-5119/2020\|EPI_ISL_690228\|2020-08 | ···················· | ···················· | ····················· | ···················· | ···················· | ···················· |
| V | 353 | hCoV-19/Japan/PG-5120/2020\|EPI_ISL_690229\|2020-08 | ···················· | ···················· | ····················· | ···················· | ···················· | ···················· |
| V | 354 | hCoV-19/Japan/PG-5121/2020\|EPI_ISL_690230\|2020-08 | ···················· | ···················· | ····················· | ···················· | ···················· | ···················· |
| V | 355 | hCoV-19/Japan/PG-5122/2020\|EPI_ISL_690231\|2020-08 | ···················· | ···················· | ····················· | ···················· | ···················· | ···················· |
| V | 356 | hCoV-19/Japan/PG-5124/2020\|EPI_ISL_690232\|2020-08 | ···················· | ···················· | ····················· | ···················· | ···················· | ···················· |
| V | 357 | hCoV-19/Japan/PG-5125/2020\|EPI_ISL_690233\|2020-08 | ···················· | ···················· | ····················· | ···················· | ···················· | ···················· |
| V | 358 | hCoV-19/Japan/PG-5126/2020\|EPI_ISL_690234\|2020-08 | ···················· | ···················· | ····················· | ···················· | ···················· | ···················· |
| V | 359 | hCoV-19/Japan/PG-5127/2020\|EPI_ISL_690235\|2020-08 | ···················· | ···················· | ····················· | ···················· | ···················· | ···················· |
| V | 360 | hCoV-19/Japan/PG-5129/2020\|EPI_ISL_690236\|2020-08 | ···················· | ···················· | ····················· | ···················· | ···················· | ···················· |
| V | 361 | hCoV-19/Japan/PG-5130/2020\|EPI_ISL_690237\|2020-08 | ···················· | ···················· | ····················· | ···················· | ···················· | ···················· |
| V | 362 | hCoV-19/Japan/PG-5131/2020\|EPI_ISL_690238\|2020-08 | ···················· | ···················· | ····················· | ···················· | ···················· | ···················· |
| V | 363 | hCoV-19/Japan/PG-5132/2020\|EPI_ISL_690239\|2020-08 | ···················· | ···················· | ····················· | ···················· | ···················· | ···················· |
| V | 364 | hCoV-19/Japan/PG-5133/2020\|EPI_ISL_690240\|2020-08 | ···················· | ···················· | ····················· | ···················· | ···················· | ···················· |
| V | 365 | hCoV-19/Japan/PG-5134/2020\|EPI_ISL_690241\|2020-08 | ···················· | ···················· | ····················· | ···················· | ···················· | ···················· |
| V | 366 | hCoV-19/Japan/PG-5135/2020\|EPI_ISL_690242\|2020-08 | ···················· | ···················· | ····················· | ···················· | ···················· | ···················· |
| V | 367 | hCoV-19/Japan/PG-5136/2020\|EPI_ISL_690243\|2020-08 | ···················· | ···················· | ····················· | ···················· | ···················· | ···················· |
| V | 368 | hCoV-19/Japan/PG-5137/2020\|EPI_ISL_690244\|2020-08 | ···················· | ···················· | ····················· | ···················· | ···················· | ···················· |
| V | 369 | hCoV-19/Japan/PG-5138/2020\|EPI_ISL_690245\|2020-08 | ···················· | ···················· | ····················· | ···················· | ···················· | ···················· |
| V | 370 | hCoV-19/Japan/PG-5139/2020\|EPI_ISL_690246\|2020-08 | ···················· | ···················· | ····················· | ···················· | ···················· | ···················· |
| V | 371 | hCoV-19/Japan/PG-5140/2020\|EPI_ISL_690247\|2020-08 | ···················· | ···················· | ····················· | ···················· | ···················· | ···················· |
| V | 372 | hCoV-19/Japan/PG-5141/2020\|EPI_ISL_690248\|2020-08 | ···················· | ···················· | ····················· | ···················· | ···················· | ···················· |
| V | 373 | hCoV-19/Japan/PG-5142/2020\|EPI_ISL_690249\|2020-08 | ···················· | ···················· | ····················· | ···················· | ···················· | ···················· |
| V | 374 | hCoV-19/Japan/PG-5143/2020\|EPI_ISL_690250\|2020-08 | ···················· | ···················· | ····················· | ···················· | ···················· | ···················· |
| V | 375 | hCoV-19/Japan/PG-5144/2020\|EPI_ISL_690251\|2020-08 | ···················· | ···················· | ····················· | ···················· | ···················· | ···················· |
| V | 377 | hCoV-19/Japan/PG-5146/2020\|EPI_ISL_690253\|2020-08 | ···················· | ···················· | ····················· | ···················· | ···················· | ···················· |
| V | 378 | hCoV-19/Japan/PG-5147/2020\|EPI_ISL_690254\|2020-08 | ···················· | ···················· | ····················· | ···················· | ···················· | ···················· |
| V | 379 | hCoV-19/Japan/PG-5148/2020\|EPI_ISL_690255\|2020-08 | ···················· | ···················· | ····················· | ···················· | ···················· | ···················· |
| V | 380 | hCoV-19/Japan/PG-5149/2020\|EPI_ISL_690256\|2020-08 | ···················· | ···················· | ····················· | ···················· | ···················· | ···················· |
| V | 381 | hCoV-19/Japan/PG-5150/2020\|EPI_ISL_690257\|2020-08 | ···················· | ···················· | ····················· | ···················· | ···················· | ···················· |
| V | 382 | hCoV-19/Japan/PG-5151/2020\|EPI_ISL_690258\|2020-08 | ···················· | ···················· | ····················· | ···················· | ···················· | ···················· |
| V | 383 | hCoV-19/Japan/PG-5152/2020\|EPI_ISL_690259\|2020-08 | ···················· | ···················· | ····················· | ···················· | ···················· | ···················· |
| V | 384 | hCoV-19/Japan/PG-5153/2020\|EPI_ISL_690260\|2020-08 | ···················· | ···················· | ····················· | ···················· | ···················· | ···················· |
| V | 386 | hCoV-19/Japan/PG-5155/2020\|EPI_ISL_690262\|2020-08 | ···················· | ···················· | ····················· | ···················· | ···················· | ···················· |
| V | 387 | hCoV-19/Japan/PG-5156/2020\|EPI_ISL_690263\|2020-08 | ···················· | ···················· | ····················· | ···················· | ···················· | ···················· |
| V | 388 | hCoV-19/Japan/PG-5157/2020\|EPI_ISL_690264\|2020-08 | ···················· | ···················· | ····················· | ···················· | ···················· | ···················· |
| V | 389 | hCoV-19/Japan/PG-5158/2020\|EPI_ISL_690265\|2020-08 | ···················· | ···················· | ····················· | ···················· | ···················· | ···················· |
| V | 390 | hCoV-19/Japan/PG-5159/2020\|EPI_ISL_690266\|2020-08 | ···················· | ···················· | ····················· | ···················· | ···················· | ···················· |
| V | 391 | hCoV-19/Japan/PG-5160/2020\|EPI_ISL_690267\|2020-08 | ···················· | ···················· | ····················· | ···················· | ···················· | ···················· |
| V | 392 | hCoV-19/Japan/PG-5162/2020\|EPI_ISL_690268\|2020-08 | ···················· | ···················· | ····················· | ···················· | ···················· | ···················· |
| V | 393 | hCoV-19/Japan/PG-5163/2020\|EPI_ISL_690269\|2020-08 | ···················· | ···················· | ····················· | ···················· | ···················· | ···················· |
| V | 394 | hCoV-19/Japan/PG-5167/2020\|EPI_ISL_690270\|2020-08 | ···················· | ···················· | ····················· | ···················· | ···················· | ···················· |
| V | 395 | hCoV-19/Japan/PG-5168/2020\|EPI_ISL_690271\|2020-08 | ···················· | ···················· | ····················· | ···················· | ···················· | ···················· |
| V | 396 | hCoV-19/Japan/PG-5169/2020\|EPI_ISL_690272\|2020-08 | ···················· | ···················· | ····················· | ···················· | ···················· | ···················· |
| V | 397 | hCoV-19/Japan/PG-5170/2020\|EPI_ISL_690273\|2020-08 | ···················· | ···················· | ····················· | ···················· | ···················· | ···················· |
| V | 398 | hCoV-19/Japan/PG-3389/2020\|EPI_ISL_691503\|2020-05 | ···················· | ···················· | ····················· | ···················· | ···················· | ···················· |
| V | 399 | hCoV-19/Japan/PG-3390/2020\|EPI_ISL_691504\|2020-05 | ···················· | ···················· | ····················· | ···················· | ···················· | ···················· |
| V | 400 | hCoV-19/Japan/PG-3391/2020\|EPI_ISL_691505\|2020-05 | ···················· | ···················· | ····················· | ···················· | ···················· | ···················· |
| V | 401 | hCoV-19/Japan/PG-3392/2020\|EPI_ISL_691506\|2020-05 | ···················· | ···················· | ····················· | ···················· | ···················· | ···················· |
| V | 402 | hCoV-19/Japan/PG-3395/2020\|EPI_ISL_691509\|2020-04 | ···················· | ···················· | ····················· | ···················· | ···················· | ···················· |
| V | 403 | hCoV-19/Japan/PG-3408/2020\|EPI_ISL_691522\|2020-04 | ···················· | ···················· | ····················· | ···················· | ···················· | ···················· |
| V | 404 | hCoV-19/Japan/PG-3418/2020\|EPI_ISL_691530\|2020-04 | ···················· | ···················· | ····················· | ···················· | ···················· | ···················· |
| V | 405 | hCoV-19/Japan/PG-3419/2020\|EPI_ISL_691531\|2020-04 | ···················· | ···················· | ····················· | ···················· | ···················· | ···················· |
| V | 406 | hCoV-19/Japan/PG-3420/2020\|EPI_ISL_691532\|2020-04 | ···················· | ···················· | ····················· | ···················· | ···················· | ···················· |
| V | 407 | hCoV-19/Japan/PG-3421/2020\|EPI_ISL_691533\|2020-04 | ···················· | ···················· | ····················· | ···················· | ···················· | ···················· |
| V | 408 | hCoV-19/Japan/PG-3422/2020\|EPI_ISL_691534\|2020-04 | ···················· | ···················· | ····················· | ···················· | ···················· | ···················· |
| V | 409 | hCoV-19/Japan/PG-3423/2020\|EPI_ISL_691535\|2020-04 | ···················· | ···················· | ····················· | ···················· | ···················· | ···················· |
| V | 412 | hCoV-19/Japan/PG-3426/2020\|EPI_ISL_691538\|2020-04 | ···················· | ···················· | ····················· | ···················· | ···················· | ···················· |
| V | 414 | hCoV-19/Japan/PG-3429/2020\|EPI_ISL_691540\|2020-04 | ···················· | ···················· | ····················· | ···················· | ···················· | ···················· |
| V | 415 | hCoV-19/Japan/PG-3430/2020\|EPI_ISL_691541\|2020-04 | ···················· | ···················· | ····················· | ···················· | ···················· | ···················· |
| V | 416 | hCoV-19/Japan/PG-3431/2020\|EPI_ISL_691542\|2020-04 | ···················· | ···················· | ····················· | ···················· | ···················· | ···················· |
| V | 417 | hCoV-19/Japan/PG-3432/2020\|EPI_ISL_691543\|2020-07 | ···················· | ···················· | ····················· | ···················· | ···················· | ···················· |
| V | 418 | hCoV-19/Japan/PG-3433/2020\|EPI_ISL_691544\|2020-07 | ···················· | ···················· | ····················· | ···················· | ···················· | ···················· |
| V | 419 | hCoV-19/Japan/PG-3434/2020\|EPI_ISL_691545\|2020-07 | ···················· | ···················· | ····················· | ···················· | ···················· | ···················· |
| V | 420 | hCoV-19/Japan/PG-3435/2020\|EPI_ISL_691546\|2020-06 | ···················· | ···················· | ····················· | ···················· | ···················· | ···················· |
| V | 421 | hCoV-19/Japan/PG-3436/2020\|EPI_ISL_691547\|2020-07 | ···················· | ···················· | ····················· | ···················· | ···················· | ···················· |
| V | 422 | hCoV-19/Japan/PG-3437/2020\|EPI_ISL_691548\|2020-06 | ···················· | ···················· | ····················· | ···················· | ···················· | ···················· |
| V | 423 | hCoV-19/Japan/PG-3439/2020\|EPI_ISL_691549\|2020-06 | ···················· | ···················· | ····················· | ···················· | ···················· | ···················· |
| V | 424 | hCoV-19/Japan/PG-3440/2020\|EPI_ISL_691550\|2020-07 | ···················· | ·A·················· | ····················· | ···················· | ···················· | ···················· |
| V | 425 | hCoV-19/Japan/PG-3441/2020\|EPI_ISL_691551\|2020-07 | ···················· | ·A·················· | ····················· | ···················· | ···················· | ···················· |
| V | 426 | hCoV-19/Japan/PG-3442/2020\|EPI_ISL_691552\|2020-06 | ···················· | ···················· | ····················· | ···················· | ···················· | ···················· |
| V | 427 | hCoV-19/Japan/PG-3443/2020\|EPI_ISL_691553\|2020-06 | ···················· | ···················· | ····················· | ···················· | ···················· | ···················· |
| V | 428 | hCoV-19/Japan/PG-3444/2020\|EPI_ISL_691554\|2020-06 | ···················· | ···················· | ····················· | ···················· | ···················· | ···················· |
| V | 429 | hCoV-19/Japan/PG-3445/2020\|EPI_ISL_691555\|2020-06 | ···················· | ···················· | ····················· | ···················· | ···················· | ···················· |
| V | 430 | hCoV-19/Japan/PG-3446/2020\|EPI_ISL_691556\|2020-07 | ···················· | ···················· | ····················· | ···················· | ···················· | ···················· |

| | | | | | | | |
|---|---|---|---|---|---|---|---|
| V | 431 hCoV-19/Japan/PG-3447/2020\|EPI_ISL_691557\|2020-07 | .................... | .................... | ..................... | .................... | .................... | .................... |
| V | 432 hCoV-19/Japan/PG-3448/2020\|EPI_ISL_691558\|2020-07 | .................... | .................... | ..................... | .................... | .................... | .................... |
| V | 433 hCoV-19/Japan/PG-3449/2020\|EPI_ISL_691559\|2020-06 | .................... | .................... | ..................... | .................... | .................... | .................... |
| V | 434 hCoV-19/Japan/PG-3450/2020\|EPI_ISL_691560\|2020-07 | .................... | .................... | ..................... | .................... | .................... | .................... |
| V | 435 hCoV-19/Japan/PG-3451/2020\|EPI_ISL_691561\|2020-07 | .................... | .................... | ..................... | .................... | .................... | .................... |
| V | 436 hCoV-19/Japan/PG-3452/2020\|EPI_ISL_691562\|2020-07 | .................... | .................... | ..................... | .................... | .................... | .................... |
| V | 437 hCoV-19/Japan/PG-3453/2020\|EPI_ISL_691563\|2020-07 | .................... | .................... | ..................... | .................... | .................... | .................... |
| V | 438 hCoV-19/Japan/PG-3454/2020\|EPI_ISL_691564\|2020-07 | .................... | .................... | ..................... | .................... | .................... | .................... |
| V | 439 hCoV-19/Japan/PG-3455/2020\|EPI_ISL_691565\|2020-07 | .................... | .................... | ..................... | .................... | .................... | .................... |
| V | 440 hCoV-19/Japan/PG-3456/2020\|EPI_ISL_691566\|2020-07 | .................... | .................... | ..................... | .................... | .................... | .................... |
| V | 441 hCoV-19/Japan/PG-3457/2020\|EPI_ISL_691567\|2020-07 | .................... | .................... | ..................... | .................... | .................... | .................... |
| V | 442 hCoV-19/Japan/PG-3458/2020\|EPI_ISL_691568\|2020-07 | .................... | .................... | ..................... | .................... | .................... | .................... |
| V | 443 hCoV-19/Japan/PG-3459/2020\|EPI_ISL_691569\|2020-07 | .................... | .................... | ..................... | .................... | .................... | .................... |
| V | 444 hCoV-19/Japan/PG-3460/2020\|EPI_ISL_691570\|2020-07 | .................... | .................... | ..................... | .................... | .................... | .................... |
| V | 445 hCoV-19/Japan/PG-3461/2020\|EPI_ISL_691571\|2020-07 | .................... | .................... | ..................... | .................... | .................... | .................... |
| V | 446 hCoV-19/Japan/PG-3462/2020\|EPI_ISL_691572\|2020-07 | .................... | .................... | ..................... | .................... | .................... | .................... |
| V | 447 hCoV-19/Japan/PG-3463/2020\|EPI_ISL_691573\|2020-07 | .................... | .................... | ..................... | .................... | .................... | .................... |
| V | 448 hCoV-19/Japan/PG-3464/2020\|EPI_ISL_691574\|2020-07 | .................... | .................... | ..................... | .................... | .................... | .................... |
| V | 449 hCoV-19/Japan/PG-3465/2020\|EPI_ISL_691575\|2020-07 | .................... | .................... | ..................... | .................... | .................... | .................... |
| V | 450 hCoV-19/Japan/PG-3466/2020\|EPI_ISL_691576\|2020-07 | .................... | .................... | ..................... | .................... | .................... | .................... |
| V | 451 hCoV-19/Japan/PG-3467/2020\|EPI_ISL_691577\|2020-07 | .................... | .................... | ..................... | .................... | .................... | .................... |
| V | 452 hCoV-19/Japan/PG-3468/2020\|EPI_ISL_691578\|2020-07 | .................... | .................... | ..................... | .................... | .................... | .................... |
| V | 453 hCoV-19/Japan/PG-3469/2020\|EPI_ISL_691579\|2020-07 | .................... | .................... | ..................... | .................... | .................... | .................... |
| V | 454 hCoV-19/Japan/PG-3470/2020\|EPI_ISL_691580\|2020-07 | .................... | .................... | ..................... | .................... | .................... | .................... |
| V | 455 hCoV-19/Japan/PG-3471/2020\|EPI_ISL_691581\|2020-07 | .................... | .................... | ..................... | .................... | .................... | .................... |
| V | 456 hCoV-19/Japan/PG-3472/2020\|EPI_ISL_691582\|2020-07 | .................... | .................... | ..................... | .................... | .................... | .................... |
| V | 457 hCoV-19/Japan/PG-3473/2020\|EPI_ISL_691583\|2020-07 | .................... | .................... | ..................... | .................... | .................... | .................... |
| V | 458 hCoV-19/Japan/PG-3474/2020\|EPI_ISL_691584\|2020-07 | .................... | .................... | ..................... | .................... | .................... | .................... |
| V | 459 hCoV-19/Japan/PG-3475/2020\|EPI_ISL_691585\|2020-07 | .................... | .................... | ..................... | .................... | .................... | .................... |
| V | 460 hCoV-19/Japan/PG-3476/2020\|EPI_ISL_691586\|2020-07 | .................... | .................... | ..................... | .................... | .................... | .................... |
| V | 461 hCoV-19/Japan/PG-3477/2020\|EPI_ISL_691587\|2020-07 | .................... | .................... | ..................... | .................... | .................... | .................... |
| V | 462 hCoV-19/Japan/PG-3478/2020\|EPI_ISL_691588\|2020-07 | .................... | .................... | ..................... | .................... | .................... | .................... |
| V | 463 hCoV-19/Japan/PG-3479/2020\|EPI_ISL_691589\|2020-07 | .................... | .................... | ..................... | .................... | .................... | .................... |
| V | 464 hCoV-19/Japan/PG-3480/2020\|EPI_ISL_691590\|2020-07 | .................... | .................... | ..................... | .................... | .................... | .................... |
| V | 465 hCoV-19/Japan/PG-3481/2020\|EPI_ISL_691591\|2020-07 | .................... | .................... | ..................... | .................... | .................... | .................... |
| V | 466 hCoV-19/Japan/PG-3482/2020\|EPI_ISL_691592\|2020-07 | .................... | .................... | ..................... | .................... | .................... | .................... |
| V | 467 hCoV-19/Japan/PG-3483/2020\|EPI_ISL_691593\|2020-07 | .................... | .................... | ..................... | .................... | .................... | .................... |
| V | 468 hCoV-19/Japan/PG-3484/2020\|EPI_ISL_691594\|2020-07 | .................... | .................... | ..................... | .................... | .................... | .................... |
| V | 469 hCoV-19/Japan/PG-3485/2020\|EPI_ISL_691595\|2020-07 | .................... | .................... | ..................... | .................... | .................... | .................... |
| V | 470 hCoV-19/Japan/PG-3486/2020\|EPI_ISL_691596\|2020-07 | .................... | .................... | ..................... | .................... | .................... | .................... |
| V | 471 hCoV-19/Japan/PG-3487/2020\|EPI_ISL_691597\|2020-07 | .................... | .................... | ..................... | .................... | .................... | .................... |
| V | 472 hCoV-19/Japan/PG-3488/2020\|EPI_ISL_691598\|2020-07 | .................... | .................... | ..................... | .................... | .................... | .................... |
| V | 473 hCoV-19/Japan/PG-3490/2020\|EPI_ISL_691599\|2020-07 | .................... | .................... | ..................... | .................... | .................... | .................... |
| V | 474 hCoV-19/USA/MA-MASPHL-00991/2020\|EPI_ISL_692801\|20 | .................... | .................... | ..................... | .................... | .................... | .................... |
| V | 475 hCoV-19/USA/MA-MASPHL-00963/2020\|EPI_ISL_692809\|20 | .................... | .................... | ..................... | .................... | .................... | .................... |
| V | 476 hCoV-19/USA/MA-MASPHL-00961/2020\|EPI_ISL_692810\|20 | .................... | .................... | ..................... | .................... | .................... | .................... |
| V | 477 hCoV-19/USA/MA-MASPHL-00960/2020\|EPI_ISL_692811\|20 | .................... | .................... | ..................... | .................... | .................... | .................... |
| V | 478 hCoV-19/USA/MA-MASPHL-00959/2020\|EPI_ISL_692812\|20 | .................... | .................... | ..................... | .................... | .................... | .................... |
| V | 479 hCoV-19/USA/MA-MASPHL-00958/2020\|EPI_ISL_692813\|20 | .................... | .................... | ..................... | .................... | .................... | .................... |
| V | 480 hCoV-19/USA/MA-MASPHL-00957/2020\|EPI_ISL_692814\|20 | .................... | .................... | ..................... | .................... | .................... | .................... |
| V | 481 hCoV-19/USA/MA-MASPHL-00956/2020\|EPI_ISL_692815\|20 | .................... | .................... | ..................... | .................... | .................... | .................... |
| V | 482 hCoV-19/USA/MA-MASPHL-00955/2020\|EPI_ISL_692816\|20 | .................... | .................... | ..................... | .................... | .................... | .................... |
| V | 483 hCoV-19/USA/MA-MASPHL-00954/2020\|EPI_ISL_692817\|20 | .................... | .................... | ..................... | .................... | .................... | .................... |
| V | 484 hCoV-19/USA/MA-MASPHL-00952/2020\|EPI_ISL_692819\|20 | .................... | .................... | ..................... | .................... | .................... | .................... |
| V | 485 hCoV-19/USA/MA-MASPHL-00942/2020\|EPI_ISL_692820\|20 | .................... | .................... | ..................... | .................... | .................... | .................... |
| V | 486 hCoV-19/USA/MA-MASPHL-00935/2020\|EPI_ISL_692822\|20 | .................... | .................... | ..................... | .................... | .................... | .................... |
| V | 487 hCoV-19/USA/MA-MASPHL-00912/2020\|EPI_ISL_692835\|20 | .................... | .................... | ..................... | .................... | .................... | .................... |
| V | 488 hCoV-19/USA/MA-MASPHL-00911/2020\|EPI_ISL_692836\|20 | .................... | .................... | ..................... | .................... | .................... | .................... |
| V | 489 hCoV-19/USA/MA-MASPHL-00910/2020\|EPI_ISL_692837\|20 | .................... | .................... | ..................... | .................... | .................... | .................... |
| V | 490 hCoV-19/USA/MA-MASPHL-00909/2020\|EPI_ISL_692838\|20 | .................... | .................... | ..................... | .................... | .................... | .................... |
| V | 491 hCoV-19/USA/MA-MASPHL-00879/2020\|EPI_ISL_692853\|20 | .................... | .................... | ..................... | .................... | .................... | .................... |
| V | 492 hCoV-19/USA/MA-MASPHL-00865/2020\|EPI_ISL_692860\|20 | .................... | .................... | ..................... | .................... | .................... | .................... |
| V | 493 hCoV-19/USA/MA-MASPHL-00838/2020\|EPI_ISL_692865\|20 | .................... | .................... | ..................... | .................... | .................... | .................... |
| V | 494 hCoV-19/USA/MA-MASPHL-00836/2020\|EPI_ISL_692867\|20 | .................... | .................... | ..................... | .................... | .................... | .................... |
| V | 495 hCoV-19/USA/MA-MASPHL-00834/2020\|EPI_ISL_692869\|20 | .................... | .................... | ..................... | .................... | .................... | .................... |
| V | 496 hCoV-19/USA/MA-MASPHL-00833/2020\|EPI_ISL_692870\|20 | .................... | .................... | ..................... | .................... | .................... | .................... |
| V | 497 hCoV-19/USA/MA-MASPHL-00832/2020\|EPI_ISL_692871\|20 | .................... | .................... | ..................... | .................... | .................... | .................... |
| V | 498 hCoV-19/USA/MA-MASPHL-00828/2020\|EPI_ISL_692873\|20 | .................... | .................... | ..................... | .................... | .................... | .................... |
| V | 499 hCoV-19/USA/MA-MASPHL-00827/2020\|EPI_ISL_692874\|20 | .................... | .................... | ..................... | .................... | .................... | .................... |
| V | 500 hCoV-19/USA/MA-MASPHL-00825/2020\|EPI_ISL_692876\|20 | .................... | .................... | ..................... | .................... | .................... | .................... |
| V | 501 hCoV-19/USA/MA-MASPHL-00823/2020\|EPI_ISL_692878\|20 | .................... | .................... | ..................... | .................... | .................... | .................... |
| V | 502 hCoV-19/USA/MA-MASPHL-00821/2020\|EPI_ISL_692880\|20 | .................... | .................... | ..................... | .................... | .................... | .................... |
| V | 503 hCoV-19/USA/MA-MASPHL-00814/2020\|EPI_ISL_692886\|20 | .................... | .................... | ..................... | .................... | .................... | .................... |
| V | 504 hCoV-19/USA/MA-MASPHL-00812/2020\|EPI_ISL_692888\|20 | .................... | .................... | ..................... | .................... | .................... | .................... |
| V | 505 hCoV-19/USA/MA-MASPHL-00810/2020\|EPI_ISL_692890\|20 | .................... | .................... | ..................... | .................... | .................... | .................... |
| V | 506 hCoV-19/USA/MA-MASPHL-00809/2020\|EPI_ISL_692891\|20 | .................... | .................... | ..................... | .................... | .................... | .................... |
| V | 507 hCoV-19/USA/MA-MASPHL-00808/2020\|EPI_ISL_692892\|20 | .................... | .................... | ..................... | .................... | .................... | .................... |
| V | 508 hCoV-19/USA/MA-MASPHL-00807/2020\|EPI_ISL_692893\|20 | .................... | .................... | ..................... | .................... | .................... | .................... |
| V | 509 hCoV-19/USA/MA-MASPHL-00806/2020\|EPI_ISL_692894\|20 | .................... | .................... | ..................... | .................... | .................... | .................... |
| V | 510 hCoV-19/USA/MA-MASPHL-00805/2020\|EPI_ISL_692895\|20 | .................... | .................... | ..................... | .................... | .................... | .................... |

| | | | | | | | |
|---|---|---|---|---|---|---|---|
| V | 511 hCoV-19/USA/MA-MASPHL-00803/2020\|EPI_ISL_692897\|20 | ···················· | ···················· | ····················· | ···················· | ···················· | ···················· |
| V | 512 hCoV-19/USA/MA-MASPHL-00801/2020\|EPI_ISL_692899\|20 | ···················· | ···················· | ····················· | ···················· | ···K················ | ···················· |
| V | 513 hCoV-19/Japan/PG-0690/2020\|EPI_ISL_684300\|2020-04 | ···················· | ···················· | ····················· | ···················· | ···················· | ···················· |
| V | 514 hCoV-19/Japan/PG-0691/2020\|EPI_ISL_684301\|2020-04 | ···················· | ···················· | ····················· | ···················· | ···················· | ···················· |
| V | 515 hCoV-19/Japan/PG-0692/2020\|EPI_ISL_684302\|2020-04 | ···················· | ···················· | ····················· | ···················· | ···················· | ···················· |
| V | 516 hCoV-19/Japan/PG-0693/2020\|EPI_ISL_684303\|2020-04 | ···················· | ···················· | ····················· | ···················· | ···················· | ···················· |
| V | 517 hCoV-19/Japan/PG-0694/2020\|EPI_ISL_684304\|2020 | ···················· | ···················· | ····················· | ···················· | ···················· | ···················· |
| V | 518 hCoV-19/Japan/PG-0695/2020\|EPI_ISL_684305\|2020-04 | ···················· | ···················· | ····················· | ···················· | ···················· | ···················· |
| V | 519 hCoV-19/Japan/PG-0696/2020\|EPI_ISL_684306\|2020-04 | ···················· | ···················· | ····················· | ···················· | ···················· | ···················· |
| V | 520 hCoV-19/Japan/PG-0697/2020\|EPI_ISL_684307\|2020-04 | ···················· | ···················· | ····················· | ···················· | ···················· | ···················· |
| V | 521 hCoV-19/Japan/PG-0698/2020\|EPI_ISL_684308\|2020-04 | ···················· | ···················· | ····················· | ···················· | ···················· | ···················· |
| V | 522 hCoV-19/Japan/PG-0699/2020\|EPI_ISL_684309\|2020-04 | ···················· | ···················· | ····················· | ···················· | ···················· | ···················· |
| V | 523 hCoV-19/Japan/PG-0700/2020\|EPI_ISL_684310\|2020-04 | ···················· | ···················· | ····················· | ···················· | ···················· | ···················· |
| V | 524 hCoV-19/Japan/PG-0701/2020\|EPI_ISL_684311\|2020-04 | ···················· | ···················· | ····················· | ···················· | ···················· | ···················· |
| V | 525 hCoV-19/Japan/PG-0769/2020\|EPI_ISL_684379\|2020-03 | ···················· | ···················· | ····················· | ···················· | ···················· | ···················· |
| V | 526 hCoV-19/Japan/PG-0771/2020\|EPI_ISL_684381\|2020-03 | ···················· | ···················· | ····················· | ···················· | ···················· | ···················· |
| V | 527 hCoV-19/Japan/PG-0772/2020\|EPI_ISL_684382\|2020-03 | ···················· | ···················· | ····················· | ···················· | ···················· | ···················· |
| V | 528 hCoV-19/Japan/PG-0773/2020\|EPI_ISL_684383\|2020-03 | ···················· | ···················· | ····················· | ···················· | ···················· | ···················· |
| V | 529 hCoV-19/Japan/PG-0774/2020\|EPI_ISL_684384\|2020-03 | ···················· | ···················· | ····················· | ···················· | ···················· | ···················· |
| V | 530 hCoV-19/Japan/PG-0775/2020\|EPI_ISL_684385\|2020-04 | ···················· | ···················· | ····················· | ···················· | ···················· | ···················· |
| V | 531 hCoV-19/Japan/PG-0776/2020\|EPI_ISL_684386\|2020-04 | ···················· | ···················· | ····················· | ···················· | ···················· | ···················· |
| V | 532 hCoV-19/Japan/PG-0777/2020\|EPI_ISL_684387\|2020-04 | ···················· | ···················· | ····················· | ···················· | ···················· | ···················· |
| V | 533 hCoV-19/Japan/PG-0778/2020\|EPI_ISL_684388\|2020-04 | ···················· | ···················· | ····················· | ···················· | ···················· | ···················· |
| V | 534 hCoV-19/Japan/PG-0779/2020\|EPI_ISL_684389\|2020-04 | ···················· | ···················· | ····················· | ···················· | ···················· | ···················· |
| V | 535 hCoV-19/Japan/PG-0780/2020\|EPI_ISL_684390\|2020-04 | ···················· | ···················· | ····················· | ···················· | ···················· | ···················· |
| V | 536 hCoV-19/Japan/PG-0781/2020\|EPI_ISL_684391\|2020-04 | ···················· | ···················· | ····················· | ···················· | ···················· | ···················· |
| V | 537 hCoV-19/Japan/PG-0782/2020\|EPI_ISL_684392\|2020-04 | ···················· | ···················· | ····················· | ···················· | ···················· | ···················· |
| V | 538 hCoV-19/Japan/PG-0783/2020\|EPI_ISL_684393\|2020-04 | ···················· | ···················· | ····················· | ···················· | ···················· | ···················· |
| V | 539 hCoV-19/Japan/PG-0784/2020\|EPI_ISL_684394\|2020-04 | ···················· | ···················· | ····················· | ···················· | ···················· | ···················· |
| V | 540 hCoV-19/Japan/PG-0785/2020\|EPI_ISL_684395\|2020-04 | ···················· | ···················· | ····················· | ···················· | ···················· | ···················· |
| V | 541 hCoV-19/Japan/PG-0786/2020\|EPI_ISL_684396\|2020-04 | ···················· | ···················· | ····················· | ···················· | ···················· | ···················· |
| V | 542 hCoV-19/Japan/PG-0787/2020\|EPI_ISL_684397\|2020-04 | ···················· | ···················· | ····················· | ···················· | ···················· | ···················· |
| V | 543 hCoV-19/Japan/PG-0788/2020\|EPI_ISL_684398\|2020-04 | ···················· | ···················· | ····················· | ···················· | ···················· | ···················· |
| V | 544 hCoV-19/Japan/PG-0789/2020\|EPI_ISL_684399\|2020-04 | ···················· | ···················· | ····················· | ···················· | ···················· | ···················· |
| V | 545 hCoV-19/Japan/PG-1995/2020\|EPI_ISL_685600\|2020-04 | ···················· | ···················· | ····················· | ···················· | ···················· | ···················· |
| V | 546 hCoV-19/Japan/PG-1996/2020\|EPI_ISL_685601\|2020-04 | ···················· | ···················· | ····················· | ···················· | ···················· | ···················· |
| V | 547 hCoV-19/Japan/PG-1997/2020\|EPI_ISL_685602\|2020-04 | ···················· | ···················· | ····················· | ···················· | ···················· | ···················· |
| V | 548 hCoV-19/Japan/PG-1998/2020\|EPI_ISL_685603\|2020-04 | ···················· | ···················· | ····················· | ···················· | ···················· | ···················· |
| V | 549 hCoV-19/Japan/PG-1999/2020\|EPI_ISL_685604\|2020-05 | ···················· | ···················· | ····················· | ···················· | ···················· | ···················· |
| V | 550 hCoV-19/Japan/PG-2000/2020\|EPI_ISL_685605\|2020-05 | ···················· | ···················· | ····················· | ···················· | ···················· | ···················· |
| V | 551 hCoV-19/Japan/PG-2001/2020\|EPI_ISL_685606\|2020-03 | ···················· | ···················· | ····················· | ···················· | ···················· | ···················· |
| V | 552 hCoV-19/Japan/PG-2002/2020\|EPI_ISL_685607\|2020-04 | ···················· | ···················· | ····················· | ···················· | ···················· | ···················· |
| V | 553 hCoV-19/Japan/PG-2003/2020\|EPI_ISL_685608\|2020-04 | ···················· | ···················· | ····················· | ···················· | ···················· | ···················· |
| V | 554 hCoV-19/Japan/PG-2004/2020\|EPI_ISL_685609\|2020-04 | ···················· | ···················· | ····················· | ···················· | ···················· | ···················· |
| V | 555 hCoV-19/Japan/PG-2005/2020\|EPI_ISL_685610\|2020-04 | ···················· | ···················· | ····················· | ···················· | ···················· | ···················· |
| V | 556 hCoV-19/Japan/PG-2006/2020\|EPI_ISL_685611\|2020-04 | ···················· | ···················· | ····················· | ···················· | ···················· | ···················· |
| V | 557 hCoV-19/Japan/PG-2007/2020\|EPI_ISL_685612\|2020-04 | ···················· | ···················· | ····················· | ···················· | ···················· | ···················· |
| V | 558 hCoV-19/Japan/PG-2008/2020\|EPI_ISL_685613\|2020-04 | ···················· | ···················· | ····················· | ···················· | ···················· | ···················· |
| V | 559 hCoV-19/Japan/PG-2009/2020\|EPI_ISL_685614\|2020-04 | ···················· | ···················· | ····················· | ···················· | ···················· | ···················· |
| V | 560 hCoV-19/Japan/PG-2010/2020\|EPI_ISL_685615\|2020-04 | ···················· | ···················· | ····················· | ···················· | ···················· | ···················· |
| V | 561 hCoV-19/Japan/PG-2011/2020\|EPI_ISL_685616\|2020-04 | ···················· | ···················· | ····················· | ···················· | ···················· | ···················· |
| V | 562 hCoV-19/Japan/PG-2012/2020\|EPI_ISL_685617\|2020-03 | ···················· | ···················· | ····················· | ···················· | ···················· | ···················· |
| V | 563 hCoV-19/Japan/PG-2013/2020\|EPI_ISL_685618\|2020-03 | ···················· | ···················· | ····················· | ···················· | ···················· | ···················· |
| V | 564 hCoV-19/Japan/PG-2014/2020\|EPI_ISL_685619\|2020-03 | ···················· | ···················· | ····················· | ···················· | ···················· | ···················· |
| V | 565 hCoV-19/Japan/PG-2015/2020\|EPI_ISL_685620\|2020-03 | ···················· | ···················· | ····················· | ···················· | ···················· | ···················· |
| V | 566 hCoV-19/Japan/PG-2016/2020\|EPI_ISL_685621\|2020-03 | ···················· | ···················· | ····················· | ···················· | ···················· | ···················· |
| V | 567 hCoV-19/Japan/PG-2017/2020\|EPI_ISL_685622\|2020-03 | ···················· | ···················· | ····················· | ···················· | ···················· | ···················· |
| V | 568 hCoV-19/Japan/PG-2018/2020\|EPI_ISL_685623\|2020-04 | ···················· | ···················· | ····················· | ···················· | ···················· | ···················· |
| V | 569 hCoV-19/Japan/PG-2019/2020\|EPI_ISL_685624\|2020-04 | ···················· | ···················· | ····················· | ···················· | ···················· | ···················· |
| V | 570 hCoV-19/Japan/PG-2020/2020\|EPI_ISL_685625\|2020-04 | ···················· | ···················· | ····················· | ···················· | ···················· | ···················· |
| V | 571 hCoV-19/Japan/PG-2021/2020\|EPI_ISL_685626\|2020-04 | ···················· | ···················· | ····················· | ···················· | ···················· | ···················· |
| V | 572 hCoV-19/Japan/PG-2022/2020\|EPI_ISL_685627\|2020-04 | ···················· | ···················· | ····················· | ···················· | ···················· | ···················· |
| V | 573 hCoV-19/Japan/PG-2023/2020\|EPI_ISL_685628\|2020-04 | ···················· | ···················· | ····················· | ···················· | ···················· | ···················· |
| V | 574 hCoV-19/Japan/PG-2024/2020\|EPI_ISL_685629\|2020-04 | ···················· | ···················· | ····················· | ···················· | ···················· | ···················· |
| V | 575 hCoV-19/Japan/PG-2025/2020\|EPI_ISL_685630\|2020-04 | ···················· | ···················· | ····················· | ···················· | ···················· | ···················· |
| V | 576 hCoV-19/Japan/PG-2026/2020\|EPI_ISL_685631\|2020-04 | ···················· | ···················· | ····················· | ···················· | ···················· | ···················· |
| V | 577 hCoV-19/Japan/PG-2027/2020\|EPI_ISL_685632\|2020-04 | ···················· | ···················· | ····················· | ···················· | ···················· | ···················· |
| V | 578 hCoV-19/Japan/PG-2028/2020\|EPI_ISL_685633\|2020-04 | ···················· | ···················· | ····················· | ···················· | ···················· | ···················· |
| V | 579 hCoV-19/Japan/PG-2029/2020\|EPI_ISL_685634\|2020-04 | ···················· | ···················· | ····················· | ···················· | ···················· | ···················· |
| V | 580 hCoV-19/Japan/PG-2030/2020\|EPI_ISL_685635\|2020-04 | ···················· | ···················· | ····················· | ···················· | ···················· | ···················· |
| V | 581 hCoV-19/Japan/PG-2031/2020\|EPI_ISL_685636\|2020-04 | ···················· | ···················· | ····················· | ···················· | ···················· | ···················· |
| V | 582 hCoV-19/Japan/PG-2032/2020\|EPI_ISL_685637\|2020-04 | ···················· | ···················· | ····················· | ···················· | ···················· | ···················· |
| V | 583 hCoV-19/Japan/PG-2033/2020\|EPI_ISL_685638\|2020-05 | ···················· | ···················· | ····················· | ···················· | ···················· | ···················· |
| V | 584 hCoV-19/Japan/PG-2034/2020\|EPI_ISL_685639\|2020-05 | ···················· | ···················· | ····················· | ···················· | ···················· | ···················· |
| V | 585 hCoV-19/Japan/PG-2035/2020\|EPI_ISL_685640\|2020-05 | ···················· | ···················· | ····················· | ···················· | ···················· | ···················· |
| V | 586 hCoV-19/Japan/PG-2036/2020\|EPI_ISL_685641\|2020-05 | ···················· | ···················· | ····················· | ···················· | ···················· | ···················· |
| V | 587 hCoV-19/Japan/PG-2037/2020\|EPI_ISL_685642\|2020-05 | ···················· | ···················· | ····················· | ···················· | ···················· | ···················· |
| V | 588 hCoV-19/Japan/PG-7798/2020\|EPI_ISL_686900\|2020-07 | ···················· | ···················· | ····················· | ···················· | ···················· | ···················· |
| V | 589 hCoV-19/Japan/PG-7799/2020\|EPI_ISL_686901\|2020-07 | ···················· | ···················· | ····················· | ···················· | ···················· | ···················· |
| V | 590 hCoV-19/Japan/PG-7800/2020\|EPI_ISL_686902\|2020-07 | ···················· | ···················· | ····················· | ···················· | ···················· | ···················· |

| | | | | | | | | |
|---|---|---|---|---|---|---|---|---|
| V | 591 | hCoV-19/Japan/PG-7801/2020\|EPI_ISL_686903\|2020-07 | .................... | .................... | ..................... | .................... | .................... | .................... |
| V | 592 | hCoV-19/Japan/PG-7802/2020\|EPI_ISL_686904\|2020-07 | .................... | .................... | ..................... | .................... | .................... | .................... |
| V | 593 | hCoV-19/Japan/PG-7803/2020\|EPI_ISL_686905\|2020-07 | .................... | .................... | ..................... | .................... | .................... | .................... |
| V | 594 | hCoV-19/Japan/PG-7804/2020\|EPI_ISL_686906\|2020-07 | .................... | .................... | ..................... | .................... | .................... | .................... |
| V | 595 | hCoV-19/Japan/PG-7805/2020\|EPI_ISL_686907\|2020-07 | .................... | .................... | ..................... | .................... | .................... | .................... |
| V | 596 | hCoV-19/Japan/PG-7806/2020\|EPI_ISL_686908\|2020-07 | .................... | .................... | ..................... | .................... | .................... | .................... |
| V | 597 | hCoV-19/Japan/PG-7807/2020\|EPI_ISL_686909\|2020-07 | .................... | .................... | ..................... | .................... | .................... | .................... |
| V | 598 | hCoV-19/Japan/PG-7808/2020\|EPI_ISL_686910\|2020-07 | .................... | .................... | ..................... | .................... | .................... | .................... |
| V | 599 | hCoV-19/Japan/PG-7809/2020\|EPI_ISL_686911\|2020-07 | .................... | .................... | ..................... | .................... | .................... | .................... |
| V | 600 | hCoV-19/Japan/PG-7810/2020\|EPI_ISL_686912\|2020-07 | .................... | .................... | ..................... | .................... | .................... | .................... |
| V | 601 | hCoV-19/Japan/PG-7811/2020\|EPI_ISL_686913\|2020-07 | .................... | .................... | ..................... | .................... | .................... | .................... |
| V | 602 | hCoV-19/Japan/PG-7812/2020\|EPI_ISL_686914\|2020-07 | .................... | .................... | ..................... | .................... | .................... | .................... |
| V | 603 | hCoV-19/Japan/PG-7813/2020\|EPI_ISL_686915\|2020-07 | .................... | .................... | ..................... | .................... | .................... | .................... |
| V | 604 | hCoV-19/Japan/PG-7814/2020\|EPI_ISL_686916\|2020-07 | .................... | .................... | ..................... | .................... | .................... | .................... |
| V | 605 | hCoV-19/Japan/PG-7815/2020\|EPI_ISL_686917\|2020-07 | .................... | .................... | ..................... | .................... | .................... | .................... |
| V | 606 | hCoV-19/Japan/PG-7816/2020\|EPI_ISL_686918\|2020-07 | .................... | .................... | ..................... | .................... | .................... | .................... |
| V | 607 | hCoV-19/Japan/PG-7817/2020\|EPI_ISL_686919\|2020-08 | .................... | .................... | ..................... | .................... | .................... | .................... |
| V | 608 | hCoV-19/Japan/PG-7818/2020\|EPI_ISL_686920\|2020-08 | .................... | .................... | ..................... | .................... | .................... | .................... |
| V | 609 | hCoV-19/Japan/PG-7819/2020\|EPI_ISL_686921\|2020-08 | .................... | .................... | ..................... | .................... | .................... | .................... |
| V | 610 | hCoV-19/Japan/PG-7820/2020\|EPI_ISL_686922\|2020-08 | .................... | .................... | ..................... | .................... | .................... | .................... |
| V | 611 | hCoV-19/Japan/PG-7821/2020\|EPI_ISL_686923\|2020-08 | .................... | .................... | ..................... | .................... | .................... | .................... |
| V | 612 | hCoV-19/Japan/PG-7822/2020\|EPI_ISL_686924\|2020-08 | .................... | .................... | ..................... | .................... | .................... | .................... |
| V | 613 | hCoV-19/Japan/PG-7823/2020\|EPI_ISL_686925\|2020-08 | .................... | .................... | ..................... | .................... | .................... | .................... |
| V | 614 | hCoV-19/Japan/PG-7824/2020\|EPI_ISL_686926\|2020-08 | .................... | .................... | ..................... | .................... | .................... | .................... |
| V | 615 | hCoV-19/Japan/PG-7825/2020\|EPI_ISL_686927\|2020-08 | .................... | .................... | ..................... | .................... | .................... | .................... |
| V | 616 | hCoV-19/Japan/PG-7826/2020\|EPI_ISL_686928\|2020-08 | .................... | .................... | ..................... | .................... | .................... | .................... |
| V | 617 | hCoV-19/Japan/PG-7827/2020\|EPI_ISL_686929\|2020-08 | .................... | .................... | ..................... | .................... | .................... | .................... |
| V | 618 | hCoV-19/Japan/PG-7828/2020\|EPI_ISL_686930\|2020-08 | .................... | .................... | ..................... | .................... | .................... | .................... |
| V | 619 | hCoV-19/Japan/PG-7829/2020\|EPI_ISL_686931\|2020-08 | .................... | .................... | ..................... | .................... | .................... | .................... |
| V | 620 | hCoV-19/Japan/PG-7830/2020\|EPI_ISL_686932\|2020-08 | .................... | .................... | ..................... | .................... | .................... | .................... |
| V | 621 | hCoV-19/Japan/PG-7831/2020\|EPI_ISL_686933\|2020-08 | .................... | .................... | ..................... | .................... | .................... | .................... |
| V | 622 | hCoV-19/Japan/PG-7832/2020\|EPI_ISL_686934\|2020-08 | .................... | .................... | ..................... | .................... | .................... | .................... |
| V | 623 | hCoV-19/Japan/PG-7833/2020\|EPI_ISL_686935\|2020-08 | .................... | .................... | ..................... | .................... | .................... | .................... |
| V | 624 | hCoV-19/Japan/PG-7834/2020\|EPI_ISL_686936\|2020-08 | .................... | .................... | ..................... | .................... | .................... | .................... |
| V | 625 | hCoV-19/Japan/PG-7835/2020\|EPI_ISL_686937\|2020-08 | .................... | .................... | ..................... | .................... | .................... | .................... |
| V | 626 | hCoV-19/Japan/PG-7836/2020\|EPI_ISL_686938\|2020-08 | .................... | .................... | ..................... | .................... | .................... | .................... |
| V | 627 | hCoV-19/Japan/PG-7837/2020\|EPI_ISL_686939\|2020-08 | .................... | .................... | ..................... | .................... | .................... | .................... |
| V | 628 | hCoV-19/Japan/PG-7838/2020\|EPI_ISL_686940\|2020-08 | .................... | .................... | ..................... | .................... | .................... | .................... |
| V | 629 | hCoV-19/Japan/PG-7840/2020\|EPI_ISL_686941\|2020-08 | .................... | .................... | ..................... | .................... | .................... | .................... |
| V | 630 | hCoV-19/Japan/PG-7841/2020\|EPI_ISL_686942\|2020-08 | .................... | .................... | ..................... | .................... | .................... | .................... |
| V | 631 | hCoV-19/Japan/PG-7842/2020\|EPI_ISL_686943\|2020-08 | .................... | .................... | ..................... | .................... | .................... | .................... |
| V | 632 | hCoV-19/Japan/PG-7843/2020\|EPI_ISL_686944\|2020-08 | .................... | .................... | ..................... | .................... | .................... | .................... |
| V | 633 | hCoV-19/Japan/PG-7844/2020\|EPI_ISL_686945\|2020-08 | .................... | .................... | ..................... | .................... | .................... | .................... |
| V | 634 | hCoV-19/Japan/PG-7845/2020\|EPI_ISL_686946\|2020-08 | .................... | .................... | ..................... | .................... | .................... | .................... |
| V | 636 | hCoV-19/Japan/PG-7847/2020\|EPI_ISL_686948\|2020-08 | .................... | .................... | ..................... | .................... | .................... | .................... |
| V | 637 | hCoV-19/Japan/PG-7848/2020\|EPI_ISL_686949\|2020-08 | .................... | .................... | ..................... | .................... | .................... | .................... |
| V | 638 | hCoV-19/Japan/PG-7849/2020\|EPI_ISL_686950\|2020-07 | .................... | .................... | ..................... | .................... | .................... | .................... |
| V | 639 | hCoV-19/Japan/PG-7850/2020\|EPI_ISL_686951\|2020-07 | .................... | .................... | ..................... | .................... | .................... | .................... |
| V | 640 | hCoV-19/Japan/PG-7851/2020\|EPI_ISL_686952\|2020-07 | .................... | .................... | ..................... | .................... | .................... | .................... |
| V | 641 | hCoV-19/Japan/PG-7852/2020\|EPI_ISL_686953\|2020-07 | .................... | .................... | ..................... | .................... | .................... | .................... |
| V | 642 | hCoV-19/Japan/PG-7853/2020\|EPI_ISL_686954\|2020-07 | .................... | .................... | ..................... | .................... | .................... | .................... |
| V | 643 | hCoV-19/Japan/PG-7854/2020\|EPI_ISL_686955\|2020-07 | .................... | .................... | ..................... | .................... | .................... | .................... |
| V | 644 | hCoV-19/Japan/PG-7855/2020\|EPI_ISL_686956\|2020-07 | .................... | .................... | ..................... | .................... | .................... | .................... |
| V | 645 | hCoV-19/Japan/PG-7856/2020\|EPI_ISL_686957\|2020-06 | .................... | .................... | ..................... | .................... | .................... | .................... |
| V | 646 | hCoV-19/Japan/PG-7857/2020\|EPI_ISL_686958\|2020-07 | .................... | .................... | ..................... | .................... | .................... | .................... |
| V | 647 | hCoV-19/Japan/PG-7858/2020\|EPI_ISL_686959\|2020-07 | .................... | .................... | ..................... | .................... | .................... | .................... |
| V | 648 | hCoV-19/Japan/PG-7859/2020\|EPI_ISL_686960\|2020-07 | .................... | .................... | ..................... | .................... | .................... | .................... |
| V | 649 | hCoV-19/Japan/PG-7860/2020\|EPI_ISL_686961\|2020-07 | .................... | .................... | ..................... | .................... | .................... | .................... |
| V | 650 | hCoV-19/Japan/PG-7861/2020\|EPI_ISL_686962\|2020-07 | .................... | .................... | ..................... | .................... | .................... | .................... |
| V | 651 | hCoV-19/Japan/PG-7862/2020\|EPI_ISL_686963\|2020-07 | .................... | .................... | ..................... | .................... | .................... | .................... |
| V | 652 | hCoV-19/Japan/PG-7863/2020\|EPI_ISL_686964\|2020-07 | .................... | .................... | ..................... | .................... | .................... | .................... |
| V | 653 | hCoV-19/Japan/PG-7864/2020\|EPI_ISL_686965\|2020-07 | .................... | .................... | ..................... | .................... | .................... | .................... |
| V | 654 | hCoV-19/Japan/PG-7865/2020\|EPI_ISL_686966\|2020-07 | .................... | .................... | ..................... | .................... | .................... | .................... |
| V | 655 | hCoV-19/Japan/PG-7866/2020\|EPI_ISL_686967\|2020-07 | .................... | .................... | ..................... | .................... | .................... | .................... |
| V | 656 | hCoV-19/Japan/PG-7867/2020\|EPI_ISL_686968\|2020-07 | .................... | .................... | ..................... | .................... | .................... | .................... |
| V | 657 | hCoV-19/Japan/PG-7868/2020\|EPI_ISL_686969\|2020-07 | .................... | .................... | ..................... | .................... | .................... | .................... |
| V | 658 | hCoV-19/Japan/PG-7869/2020\|EPI_ISL_686970\|2020-07 | .................... | .................... | ..................... | .................... | .................... | .................... |
| V | 659 | hCoV-19/Japan/PG-7870/2020\|EPI_ISL_686971\|2020-07 | .................... | .................... | ..................... | .................... | .................... | .................... |
| V | 660 | hCoV-19/Japan/PG-7871/2020\|EPI_ISL_686972\|2020-07 | .................... | .................... | ..................... | .................... | .................... | .................... |
| V | 661 | hCoV-19/Japan/PG-7872/2020\|EPI_ISL_686973\|2020-07 | .................... | .................... | ..................... | .................... | .................... | .................... |
| V | 662 | hCoV-19/Japan/PG-6058/2020\|EPI_ISL_689170\|2020-07 | .................... | .................... | ..................... | .................... | .................... | .................... |
| V | 663 | hCoV-19/Japan/PG-6059/2020\|EPI_ISL_689171\|2020-07 | .................... | .................... | ..................... | .................... | .................... | .................... |
| V | 664 | hCoV-19/Japan/PG-6060/2020\|EPI_ISL_689172\|2020-07 | .................... | .................... | ..................... | .................... | .................... | .................... |
| V | 665 | hCoV-19/Japan/PG-6061/2020\|EPI_ISL_689173\|2020-07 | .................... | .................... | ..................... | .................... | .................... | .................... |
| V | 666 | hCoV-19/Japan/PG-6062/2020\|EPI_ISL_689174\|2020-07 | .................... | .................... | ..................... | .................... | .................... | .................... |
| V | 667 | hCoV-19/Japan/PG-6063/2020\|EPI_ISL_689175\|2020-07 | .................... | .................... | ..................... | .................... | .................... | .................... |
| V | 668 | hCoV-19/Japan/PG-6064/2020\|EPI_ISL_689176\|2020-07 | .................... | .................... | ..................... | .................... | .................... | .................... |
| V | 669 | hCoV-19/Japan/PG-6065/2020\|EPI_ISL_689177\|2020-07 | .................... | .................... | ..................... | .................... | .................... | .................... |
| V | 670 | hCoV-19/Japan/PG-6066/2020\|EPI_ISL_689178\|2020-07 | .................... | .................... | ..................... | .................... | .................... | .................... |
| V | 671 | hCoV-19/Japan/PG-6067/2020\|EPI_ISL_689179\|2020-07 | .................... | .................... | ..................... | .................... | .................... | .................... |

| | | | | | | | | |
|---|---|---|---|---|---|---|---|---|
| V | 672 | hCoV-19/Japan/PG-6068/2020\|EPI_ISL_689180\|2020-07 | .................... | .................... | ..................... | .................... | .................... | .................... |
| V | 673 | hCoV-19/Japan/PG-6069/2020\|EPI_ISL_689181\|2020-07 | .................... | .................... | ..................... | .................... | .................... | .................... |
| V | 674 | hCoV-19/Japan/PG-6070/2020\|EPI_ISL_689182\|2020-07 | .................... | .................... | ..................... | .................... | .................... | .................... |
| V | 675 | hCoV-19/Japan/PG-6071/2020\|EPI_ISL_689183\|2020-07 | .................... | .................... | ..................... | .................... | .................... | .................... |
| V | 676 | hCoV-19/Japan/PG-6072/2020\|EPI_ISL_689184\|2020-07 | .................... | .................... | ..................... | .................... | .................... | .................... |
| V | 677 | hCoV-19/Japan/PG-6073/2020\|EPI_ISL_689185\|2020-07 | .................... | .................... | ..................... | .................... | .................... | .................... |
| V | 678 | hCoV-19/Japan/PG-6074/2020\|EPI_ISL_689186\|2020-07 | .................... | .................... | ..................... | .................... | .................... | .................... |
| V | 679 | hCoV-19/Japan/PG-6075/2020\|EPI_ISL_689187\|2020-07 | .................... | .................... | ..................... | .................... | .................... | .................... |
| V | 680 | hCoV-19/Japan/PG-6076/2020\|EPI_ISL_689188\|2020-07 | .................... | .................... | ..................... | .................... | .................... | .................... |
| V | 681 | hCoV-19/Japan/PG-6077/2020\|EPI_ISL_689189\|2020-07 | .................... | .................... | ..................... | .................... | .................... | .................... |
| V | 682 | hCoV-19/Japan/PG-6078/2020\|EPI_ISL_689190\|2020-07 | .................... | .................... | ..................... | .................... | .................... | .................... |
| V | 683 | hCoV-19/Japan/PG-6079/2020\|EPI_ISL_689191\|2020-07 | .................... | .................... | ..................... | .................... | .................... | .................... |
| V | 684 | hCoV-19/Japan/PG-6080/2020\|EPI_ISL_689192\|2020-07 | .................... | .................... | ..................... | .................... | .................... | .................... |
| V | 685 | hCoV-19/Japan/PG-6081/2020\|EPI_ISL_689193\|2020-07 | .................... | .................... | ..................... | .................... | .................... | .................... |
| V | 686 | hCoV-19/Japan/PG-6082/2020\|EPI_ISL_689194\|2020-07 | .................... | .................... | ..................... | .................... | .................... | .................... |
| V | 687 | hCoV-19/Japan/PG-6083/2020\|EPI_ISL_689195\|2020-07 | .................... | .................... | ..................... | .................... | .................... | .................... |
| V | 688 | hCoV-19/Japan/PG-6084/2020\|EPI_ISL_689196\|2020-07 | .................... | .................... | ..................... | .................... | .................... | .................... |
| V | 689 | hCoV-19/Japan/PG-6085/2020\|EPI_ISL_689197\|2020-07 | .................... | .................... | ..................... | .................... | .................... | .................... |
| V | 690 | hCoV-19/Japan/PG-6086/2020\|EPI_ISL_689198\|2020-07 | .................... | .................... | ..................... | .................... | .................... | .................... |
| V | 691 | hCoV-19/Japan/PG-6087/2020\|EPI_ISL_689199\|2020-07 | .................... | .................... | ..................... | .................... | .................... | .................... |
| V | 692 | hCoV-19/Japan/PG-6095/2020\|EPI_ISL_689207\|2020-08 | .................... | .................... | ..................... | .................... | .................... | .................... |
| V | 693 | hCoV-19/Japan/PG-6096/2020\|EPI_ISL_689208\|2020-08 | .................... | .................... | ..................... | .................... | .................... | .................... |
| V | 694 | hCoV-19/Japan/PG-6097/2020\|EPI_ISL_689209\|2020-08 | .................... | .................... | ..................... | .................... | .................... | .................... |
| V | 695 | hCoV-19/Japan/PG-6098/2020\|EPI_ISL_689210\|2020-08 | .................... | .................... | ..................... | .................... | .................... | .................... |
| V | 696 | hCoV-19/Japan/PG-6099/2020\|EPI_ISL_689211\|2020-08 | .................... | .................... | ..................... | .................... | .................... | .................... |
| V | 697 | hCoV-19/Japan/PG-6100/2020\|EPI_ISL_689212\|2020-08 | .................... | .................... | ..................... | .................... | .................... | .................... |
| V | 698 | hCoV-19/Japan/PG-6101/2020\|EPI_ISL_689213\|2020-08 | .................... | .................... | ..................... | .................... | .................... | .................... |
| V | 699 | hCoV-19/Japan/PG-6102/2020\|EPI_ISL_689214\|2020-08 | .................... | .................... | ..................... | .................... | .................... | .................... |
| V | 700 | hCoV-19/Japan/PG-6103/2020\|EPI_ISL_689215\|2020-08 | .................... | .................... | ..................... | .................... | .................... | .................... |
| V | 701 | hCoV-19/Japan/PG-6104/2020\|EPI_ISL_689216\|2020-08 | .................... | .................... | ..................... | .................... | .................... | .................... |
| V | 702 | hCoV-19/Japan/PG-6105/2020\|EPI_ISL_689217\|2020-08 | .................... | .................... | ..................... | .................... | .................... | .................... |
| V | 703 | hCoV-19/Japan/PG-6106/2020\|EPI_ISL_689218\|2020-08 | .................... | .................... | ..................... | .................... | .................... | .................... |
| V | 704 | hCoV-19/Japan/PG-6107/2020\|EPI_ISL_689219\|2020-08 | .................... | .................... | ..................... | .................... | .................... | .................... |
| V | 705 | hCoV-19/Japan/PG-6108/2020\|EPI_ISL_689220\|2020-08 | .................... | .................... | ..................... | .................... | .................... | .................... |
| V | 706 | hCoV-19/Japan/PG-6109/2020\|EPI_ISL_689221\|2020-08 | .................... | .................... | ..................... | .................... | .................... | .................... |
| V | 707 | hCoV-19/Japan/PG-6110/2020\|EPI_ISL_689222\|2020-08 | .................... | .................... | ..................... | .................... | .................... | .................... |
| V | 708 | hCoV-19/Japan/PG-5042/2020\|EPI_ISL_690153\|2020-07 | .................... | .................... | ..................... | .................... | .................... | .................... |
| V | 709 | hCoV-19/Japan/PG-5043/2020\|EPI_ISL_690154\|2020-07 | .................... | .................... | ..................... | .................... | .................... | .................... |
| V | 710 | hCoV-19/Japan/PG-5044/2020\|EPI_ISL_690155\|2020-07 | .................... | .................... | ..................... | .................... | .................... | .................... |
| V | 711 | hCoV-19/Japan/PG-5045/2020\|EPI_ISL_690156\|2020-07 | .................... | .................... | ..................... | .................... | .................... | .................... |
| V | 712 | hCoV-19/Japan/PG-5046/2020\|EPI_ISL_690157\|2020-07 | .................... | .................... | ..................... | .................... | .................... | .................... |
| V | 713 | hCoV-19/Japan/PG-5047/2020\|EPI_ISL_690158\|2020-07 | .................... | .................... | ..................... | .................... | .................... | .................... |
| V | 714 | hCoV-19/Japan/PG-5048/2020\|EPI_ISL_690159\|2020-08 | .................... | .................... | ..................... | .................... | .................... | .................... |
| V | 715 | hCoV-19/Japan/PG-5049/2020\|EPI_ISL_690160\|2020-07 | .................... | .................... | ..................... | .................... | .................... | .................... |
| V | 716 | hCoV-19/Japan/PG-5050/2020\|EPI_ISL_690161\|2020-08 | .................... | .................... | ..................... | .................... | .................... | .................... |
| V | 717 | hCoV-19/Japan/PG-5051/2020\|EPI_ISL_690162\|2020-08 | .................... | .................... | ..................... | .................... | .................... | .................... |
| V | 718 | hCoV-19/Japan/PG-5052/2020\|EPI_ISL_690163\|2020-08 | .................... | .................... | ..................... | .................... | .................... | .................... |
| V | 719 | hCoV-19/Japan/PG-5053/2020\|EPI_ISL_690164\|2020-08 | .................... | .................... | ..................... | .................... | .................... | .................... |
| V | 720 | hCoV-19/Japan/PG-5054/2020\|EPI_ISL_690165\|2020-08 | .................... | .................... | ..................... | .................... | .................... | .................... |
| V | 721 | hCoV-19/Japan/PG-5055/2020\|EPI_ISL_690166\|2020-08 | .................... | .................... | ..................... | .................... | .................... | .................... |
| V | 722 | hCoV-19/Japan/PG-5056/2020\|EPI_ISL_690167\|2020-08 | .................... | .................... | ..................... | .................... | .................... | .................... |
| V | 723 | hCoV-19/Japan/PG-5057/2020\|EPI_ISL_690168\|2020-08 | .................... | .................... | ..................... | .................... | .................... | .................... |
| V | 724 | hCoV-19/Japan/PG-5058/2020\|EPI_ISL_690169\|2020-08 | .................... | .................... | ..................... | .................... | .................... | .................... |
| V | 725 | hCoV-19/Japan/PG-5059/2020\|EPI_ISL_690170\|2020-08 | .................... | .................... | ..................... | .................... | .................... | .................... |
| V | 726 | hCoV-19/Japan/PG-5060/2020\|EPI_ISL_690171\|2020-08 | .................... | .................... | ..................... | .................... | .................... | .................... |
| V | 727 | hCoV-19/Japan/PG-5061/2020\|EPI_ISL_690172\|2020-08 | .................... | .................... | ..................... | .................... | .................... | .................... |
| V | 728 | hCoV-19/Japan/PG-5062/2020\|EPI_ISL_690173\|2020-08 | .................... | .................... | ..................... | .................... | .................... | .................... |
| V | 729 | hCoV-19/Japan/PG-5064/2020\|EPI_ISL_690174\|2020-08 | .................... | .................... | ..................... | .................... | .................... | .................... |
| V | 730 | hCoV-19/Japan/PG-3369/2020\|EPI_ISL_691484\|2020-04 | .................... | .................... | ..................... | .................... | .................... | .................... |
| V | 731 | hCoV-19/Japan/PG-3370/2020\|EPI_ISL_691485\|2020-04 | .................... | .................... | ..................... | .................... | .................... | .................... |
| V | 732 | hCoV-19/Japan/PG-3371/2020\|EPI_ISL_691486\|2020-04 | .................... | .................... | ..................... | .................... | .................... | .................... |
| V | 733 | hCoV-19/Japan/PG-3372/2020\|EPI_ISL_691487\|2020-04 | .................... | .................... | ..................... | .................... | .................... | .................... |
| V | 734 | hCoV-19/Japan/PG-3373/2020\|EPI_ISL_691488\|2020-04 | .................... | .................... | ..................... | .................... | .................... | .................... |
| V | 735 | hCoV-19/Japan/PG-3374/2020\|EPI_ISL_691489\|2020-04 | .................... | .................... | ..................... | .................... | .................... | .................... |
| V | 736 | hCoV-19/Japan/PG-3375/2020\|EPI_ISL_691490\|2020-04 | .................... | .................... | ..................... | .................... | .................... | .................... |
| V | 737 | hCoV-19/Japan/PG-3376/2020\|EPI_ISL_691491\|2020-04 | .................... | .................... | ..................... | .................... | .................... | .................... |
| V | 738 | hCoV-19/Japan/PG-3377/2020\|EPI_ISL_691492\|2020-04 | .................... | .................... | ..................... | .................... | .................... | .................... |
| V | 739 | hCoV-19/Japan/PG-3378/2020\|EPI_ISL_691493\|2020-04 | .................... | .................... | ..................... | .................... | .................... | .................... |
| V | 740 | hCoV-19/Japan/PG-3379/2020\|EPI_ISL_691494\|2020-04 | .................... | .................... | ..................... | .................... | .................... | .................... |
| V | 741 | hCoV-19/Japan/PG-3380/2020\|EPI_ISL_691495\|2020-04 | .................... | .................... | ..................... | .................... | .................... | .................... |
| V | 742 | hCoV-19/Japan/PG-3381/2020\|EPI_ISL_691496\|2020-04 | .................... | .................... | ..................... | .................... | .................... | .................... |
| V | 744 | hCoV-19/Japan/PG-3383/2020\|EPI_ISL_691498\|2020-04 | .................... | .................... | ..................... | .................... | .................... | .................... |
| V | 746 | hCoV-19/Japan/PG-3385/2020\|EPI_ISL_691500\|2020-05 | .................... | .................... | ..................... | .................... | .................... | .................... |
| V | 747 | hCoV-19/Japan/PG-3386/2020\|EPI_ISL_691501\|2020-05 | .................... | .................... | ..................... | .................... | .................... | .................... |
| V | 748 | hCoV-19/Japan/PG-3388/2020\|EPI_ISL_691502\|2020-05 | .................... | .................... | ..................... | .................... | .................... | .................... |
| V | 749 | hCoV-19/Japan/PG-3393/2020\|EPI_ISL_691507\|2020-04 | .................... | .................... | ..................... | .................... | .................... | .................... |
| V | 750 | hCoV-19/Japan/PG-3394/2020\|EPI_ISL_691508\|2020-04 | .................... | .................... | ..................... | .................... | .................... | .................... |
| V | 751 | hCoV-19/Japan/PG-3396/2020\|EPI_ISL_691510\|2020-04 | .................... | .................... | ..................... | .................... | .................... | .................... |
| V | 752 | hCoV-19/Japan/PG-3397/2020\|EPI_ISL_691511\|2020-04 | .................... | .................... | ..................... | .................... | .................... | .................... |
| V | 753 | hCoV-19/Japan/PG-3398/2020\|EPI_ISL_691512\|2020-04 | .................... | .................... | ..................... | .................... | .................... | .................... |

| | | | | | | | |
|---|---|---|---|---|---|---|---|
| V | 754 hCoV-19/Japan/PG-3399/2020\|EPI_ISL_691513\|2020-04 | .................... | .................... | ..................... | .................... | .................... | .................... |
| V | 755 hCoV-19/Japan/PG-3400/2020\|EPI_ISL_691514\|2020-04 | .................... | .................... | ..................... | .................... | .................... | .................... |
| V | 756 hCoV-19/Japan/PG-3401/2020\|EPI_ISL_691515\|2020-04 | .................... | .................... | ..................... | .................... | .................... | .................... |
| V | 757 hCoV-19/Japan/PG-3402/2020\|EPI_ISL_691516\|2020-04 | .................... | .................... | ..................... | .................... | .................... | .................... |
| V | 758 hCoV-19/Japan/PG-3403/2020\|EPI_ISL_691517\|2020-04 | .................... | .................... | ..................... | .................... | .................... | .................... |
| V | 759 hCoV-19/Japan/PG-3404/2020\|EPI_ISL_691518\|2020-04 | .................... | .................... | ..................... | .................... | .................... | .................... |
| V | 760 hCoV-19/Japan/PG-3405/2020\|EPI_ISL_691519\|2020-04 | .................... | .................... | ..................... | .................... | .................... | .................... |
| V | 761 hCoV-19/USA/MA-MASPHL-00919/2020\|EPI_ISL_692831\|20 | .................... | .................... | ..................... | .................... | .................... | .................... |
| V | 762 hCoV-19/USA/MA-MASPHL-00918/2020\|EPI_ISL_692832\|20 | .................... | .................... | ..................... | .................... | .................... | .................... |
| V | 763 hCoV-19/USA/MA-MASPHL-00908/2020\|EPI_ISL_692839\|20 | .................... | .................... | ..................... | .................... | .................... | .................... |
| V | 764 hCoV-19/USA/MA-MASPHL-00905/2020\|EPI_ISL_692841\|20 | .................... | .................... | ..................... | .................... | .................... | .................... |
| V | 765 hCoV-19/USA/MA-MASPHL-00904/2020\|EPI_ISL_692842\|20 | .................... | .................... | ..................... | .................... | .................... | .................... |
| V | 766 hCoV-19/USA/MA-MASPHL-00887/2020\|EPI_ISL_692846\|20 | .................... | .................... | ..................... | .................... | .................... | .................... |
| V | 767 hCoV-19/USA/MA-MASPHL-00886/2020\|EPI_ISL_692847\|20 | .................... | .................... | ..................... | .................... | .................... | .................... |
| V | 768 hCoV-19/USA/MA-MASPHL-00885/2020\|EPI_ISL_692848\|20 | .................... | .................... | ..................... | .................... | .................... | .................... |
| V | 769 hCoV-19/Japan/PG-0702/2020\|EPI_ISL_684312\|2020-04 | .................... | .................... | ..................... | .................... | .................... | .................... |
| V | 770 hCoV-19/Japan/PG-0703/2020\|EPI_ISL_684313\|2020-04 | .................... | .................... | ..................... | .................... | .................... | .................... |
| V | 771 hCoV-19/Japan/PG-0704/2020\|EPI_ISL_684314\|2020-04 | .................... | .................... | ..................... | .................... | .................... | .................... |
| V | 772 hCoV-19/Japan/PG-0705/2020\|EPI_ISL_684315\|2020-04 | .................... | .................... | ..................... | .................... | .................... | .................... |
| V | 773 hCoV-19/Japan/PG-0706/2020\|EPI_ISL_684316\|2020-04 | .................... | .................... | ..................... | .................... | .................... | .................... |
| V | 774 hCoV-19/Japan/PG-0707/2020\|EPI_ISL_684317\|2020-04 | .................... | .................... | ..................... | .................... | .................... | .................... |
| V | 775 hCoV-19/Japan/PG-0708/2020\|EPI_ISL_684318\|2020-04 | .................... | .................... | ..................... | .................... | .................... | .................... |
| V | 776 hCoV-19/Japan/PG-0709/2020\|EPI_ISL_684319\|2020-04 | .................... | .................... | ..................... | .................... | .................... | .................... |
| V | 777 hCoV-19/Japan/PG-0710/2020\|EPI_ISL_684320\|2020-04 | .................... | .................... | ..................... | .................... | .................... | .................... |
| V | 778 hCoV-19/Japan/PG-0711/2020\|EPI_ISL_684321\|2020-04 | .................... | .................... | ..................... | .................... | .................... | .................... |
| V | 779 hCoV-19/Japan/PG-0712/2020\|EPI_ISL_684322\|2020-04 | .................... | .................... | ..................... | .................... | .................... | .................... |
| V | 780 hCoV-19/Japan/PG-0713/2020\|EPI_ISL_684323\|2020-04 | .................... | .................... | ..................... | .................... | .................... | .................... |
| V | 781 hCoV-19/Japan/PG-0714/2020\|EPI_ISL_684324\|2020-04 | .................... | .................... | ..................... | .................... | .................... | .................... |
| V | 782 hCoV-19/Japan/PG-0715/2020\|EPI_ISL_684325\|2020-04 | .................... | .................... | ..................... | .................... | .................... | .................... |
| V | 783 hCoV-19/Japan/PG-0716/2020\|EPI_ISL_684326\|2020-04 | .................... | .................... | ..................... | .................... | .................... | .................... |
| V | 784 hCoV-19/Japan/PG-0717/2020\|EPI_ISL_684327\|2020-04 | .................... | .................... | ..................... | .................... | .................... | .................... |
| V | 785 hCoV-19/Japan/PG-0718/2020\|EPI_ISL_684328\|2020-04 | .................... | .................... | ..................... | .................... | .................... | .................... |
| V | 787 hCoV-19/Japan/PG-0720/2020\|EPI_ISL_684330\|2020-04 | .................... | .................... | ..................... | .................... | .................... | .................... |
| V | 788 hCoV-19/Japan/PG-0721/2020\|EPI_ISL_684331\|2020-04 | .................... | .................... | ..................... | .................... | .................... | .................... |
| V | 789 hCoV-19/Japan/PG-0722/2020\|EPI_ISL_684332\|2020-04 | .................... | .................... | ..................... | .................... | .................... | .................... |
| V | 790 hCoV-19/Japan/PG-0723/2020\|EPI_ISL_684333\|2020-04 | .................... | .................... | ..................... | .................... | .................... | .................... |
| V | 791 hCoV-19/Japan/PG-0724/2020\|EPI_ISL_684334\|2020-04 | .................... | .................... | ..................... | .................... | .................... | .................... |
| V | 792 hCoV-19/Japan/PG-0725/2020\|EPI_ISL_684335\|2020-04 | .................... | .................... | ..................... | .................... | .................... | .................... |
| V | 793 hCoV-19/Japan/PG-0726/2020\|EPI_ISL_684336\|2020-04 | .................... | .................... | ..................... | .................... | .................... | .................... |
| V | 794 hCoV-19/Japan/PG-0727/2020\|EPI_ISL_684337\|2020-04 | .................... | .................... | ..................... | .................... | .................... | .................... |
| V | 795 hCoV-19/Japan/PG-0728/2020\|EPI_ISL_684338\|2020-04 | .................... | .................... | ..................... | .................... | .................... | .................... |
| V | 796 hCoV-19/Japan/PG-0729/2020\|EPI_ISL_684339\|2020-04 | .................... | .................... | ..................... | .................... | .................... | .................... |
| V | 797 hCoV-19/Japan/PG-0730/2020\|EPI_ISL_684340\|2020-04 | .................... | .................... | ..................... | .................... | .................... | .................... |
| V | 798 hCoV-19/Japan/PG-0731/2020\|EPI_ISL_684341\|2020-04 | .................... | .................... | ..................... | .................... | .................... | .................... |
| V | 799 hCoV-19/Japan/PG-0732/2020\|EPI_ISL_684342\|2020-04 | .................... | .................... | ..................... | .................... | .................... | .................... |
| V | 800 hCoV-19/Japan/PG-0733/2020\|EPI_ISL_684343\|2020-04 | .................... | .................... | ..................... | .................... | .................... | .................... |
| V | 801 hCoV-19/Japan/PG-0734/2020\|EPI_ISL_684344\|2020-04 | .................... | .................... | ..................... | .................... | .................... | .................... |
| V | 802 hCoV-19/Japan/PG-0735/2020\|EPI_ISL_684345\|2020-04 | .................... | .................... | ..................... | .................... | .................... | .................... |
| V | 803 hCoV-19/Japan/PG-0736/2020\|EPI_ISL_684346\|2020-04 | .................... | .................... | ..................... | .................... | .................... | .................... |
| V | 804 hCoV-19/Japan/PG-0737/2020\|EPI_ISL_684347\|2020-04 | .................... | .................... | ..................... | .................... | .................... | .................... |
| V | 805 hCoV-19/Japan/PG-0738/2020\|EPI_ISL_684348\|2020-04 | .................... | .................... | ..................... | .................... | .................... | .................... |
| V | 806 hCoV-19/Japan/PG-0739/2020\|EPI_ISL_684349\|2020-04 | .................... | .................... | ..................... | .................... | .................... | .................... |
| V | 807 hCoV-19/Japan/PG-0740/2020\|EPI_ISL_684350\|2020-04 | .................... | .................... | ..................... | .................... | .................... | .................... |
| V | 808 hCoV-19/Japan/PG-0741/2020\|EPI_ISL_684351\|2020-04 | .................... | .................... | ..................... | .................... | .................... | .................... |
| V | 809 hCoV-19/Japan/PG-0742/2020\|EPI_ISL_684352\|2020-04 | .................... | .................... | ..................... | .................... | .................... | .................... |
| V | 810 hCoV-19/Japan/PG-0743/2020\|EPI_ISL_684353\|2020-04 | .................... | .................... | ..................... | .................... | .................... | .................... |
| V | 811 hCoV-19/Japan/PG-0744/2020\|EPI_ISL_684354\|2020-04 | .................... | .................... | ..................... | .................... | .................... | .................... |
| V | 812 hCoV-19/Japan/PG-0745/2020\|EPI_ISL_684355\|2020-04 | .................... | .................... | ..................... | .................... | .................... | .................... |
| V | 813 hCoV-19/Japan/PG-0746/2020\|EPI_ISL_684356\|2020-04 | .................... | .................... | ..................... | .................... | .................... | .................... |
| V | 814 hCoV-19/Japan/PG-0747/2020\|EPI_ISL_684357\|2020-04 | .................... | .................... | ..................... | .................... | .................... | .................... |
| V | 815 hCoV-19/Japan/PG-0748/2020\|EPI_ISL_684358\|2020-04 | .................... | .................... | ..................... | .................... | .................... | .................... |
| V | 816 hCoV-19/Japan/PG-0749/2020\|EPI_ISL_684359\|2020-04 | .................... | .................... | ..................... | .................... | .................... | .................... |
| V | 817 hCoV-19/Japan/PG-0750/2020\|EPI_ISL_684360\|2020-04 | .................... | .................... | ..................... | .................... | .................... | .................... |
| V | 818 hCoV-19/Japan/PG-0751/2020\|EPI_ISL_684361\|2020-03 | .................... | .................... | ..................... | .................... | .................... | .................... |
| V | 819 hCoV-19/Japan/PG-0752/2020\|EPI_ISL_684362\|2020-04 | .................... | .................... | ..................... | .................... | .................... | .................... |
| V | 820 hCoV-19/Japan/PG-0753/2020\|EPI_ISL_684363\|2020-04 | .................... | .................... | ..................... | .................... | .................... | .................... |
| V | 821 hCoV-19/Japan/PG-0755/2020\|EPI_ISL_684365\|2020-04 | .................... | .................... | ..................... | .................... | .................... | .................... |
| V | 822 hCoV-19/Japan/PG-0756/2020\|EPI_ISL_684366\|2020-04 | .................... | .................... | ..................... | .................... | .................... | .................... |
| V | 823 hCoV-19/Japan/PG-0757/2020\|EPI_ISL_684367\|2020-04 | .................... | .................... | ..................... | .................... | .................... | .................... |
| V | 824 hCoV-19/Japan/PG-0758/2020\|EPI_ISL_684368\|2020-04 | .................... | .................... | ..................... | .................... | .................... | .................... |
| V | 825 hCoV-19/Japan/PG-0759/2020\|EPI_ISL_684369\|2020-04 | .................... | .................... | ..................... | .................... | .................... | .................... |
| V | 826 hCoV-19/Japan/PG-0760/2020\|EPI_ISL_684370\|2020-03 | .................... | .................... | ..................... | .................... | .................... | .................... |
| V | 827 hCoV-19/Japan/PG-0761/2020\|EPI_ISL_684371\|2020-03 | .................... | .................... | ..................... | .................... | .................... | .................... |
| V | 828 hCoV-19/Japan/PG-0762/2020\|EPI_ISL_684372\|2020-03 | .................... | .................... | ..................... | .................... | .................... | .................... |
| V | 829 hCoV-19/Japan/PG-0763/2020\|EPI_ISL_684373\|2020-03 | .................... | .................... | ..................... | .................... | .................... | .................... |
| V | 830 hCoV-19/Japan/PG-0764/2020\|EPI_ISL_684374\|2020-03 | .................... | .................... | ..................... | .................... | .................... | .................... |
| V | 831 hCoV-19/Japan/PG-0765/2020\|EPI_ISL_684375\|2020-03 | .................... | .................... | ..................... | .................... | .................... | .................... |
| V | 832 hCoV-19/Japan/PG-0766/2020\|EPI_ISL_684376\|2020-03 | .................... | .................... | ..................... | .................... | .................... | .................... |
| V | 833 hCoV-19/Japan/PG-0767/2020\|EPI_ISL_684377\|2020-03 | .................... | .................... | ..................... | .................... | .................... | .................... |
| V | 834 hCoV-19/Japan/PG-0768/2020\|EPI_ISL_684378\|2020-03 | .................... | .................... | ..................... | .................... | .................... | .................... |

| | | | | | | | |
|---|---|---|---|---|---|---|---|
| V | 835 hCoV-19/Japan/PG-2038/2020\|EPI_ISL_685643\|2020-05 | .................... | .................... | ..................... | .................... | .................... | .................... |
| V | 837 hCoV-19/Japan/PG-2040/2020\|EPI_ISL_685645\|2020-05 | .................... | .................... | ..................... | .................... | .................... | .................... |
| V | 838 hCoV-19/Japan/PG-2041/2020\|EPI_ISL_685646\|2020-05 | .................... | .................... | ..................... | .................... | .................... | .................... |
| V | 839 hCoV-19/Japan/PG-2042/2020\|EPI_ISL_685647\|2020-03 | .................... | .................... | ..................... | .................... | .................... | .................... |
| V | 840 hCoV-19/Japan/PG-2043/2020\|EPI_ISL_685648\|2020-03 | .................... | .................... | ..................... | .................... | .................... | .................... |
| V | 841 hCoV-19/Japan/PG-2044/2020\|EPI_ISL_685649\|2020-04 | .................... | .................... | ..................... | .................... | .................... | .................... |
| V | 842 hCoV-19/Japan/PG-2045/2020\|EPI_ISL_685650\|2020-04 | .................... | .................... | ..................... | .................... | .................... | .................... |
| V | 843 hCoV-19/Japan/PG-2046/2020\|EPI_ISL_685651\|2020-04 | .................... | .................... | ..................... | .................... | .................... | .................... |
| V | 844 hCoV-19/Japan/PG-2047/2020\|EPI_ISL_685652\|2020-04 | .................... | .................... | ..................... | .................... | .................... | .................... |
| V | 845 hCoV-19/Japan/PG-2048/2020\|EPI_ISL_685653\|2020-04 | .................... | .................... | ..................... | .................... | .................... | .................... |
| V | 846 hCoV-19/Japan/PG-2049/2020\|EPI_ISL_685654\|2020-04 | .................... | .................... | ..................... | .................... | .................... | .................... |
| V | 847 hCoV-19/Japan/PG-2050/2020\|EPI_ISL_685655\|2020-04 | .................... | .................... | ..................... | .................... | .................... | .................... |
| V | 848 hCoV-19/Japan/PG-2051/2020\|EPI_ISL_685656\|2020-04 | .................... | .................... | ..................... | .................... | .................... | .................... |
| V | 849 hCoV-19/Japan/PG-2052/2020\|EPI_ISL_685657\|2020-04 | .................... | .................... | ..................... | .................... | .................... | .................... |
| V | 850 hCoV-19/Japan/PG-2053/2020\|EPI_ISL_685658\|2020-04 | .................... | .................... | ..................... | .................... | .................... | .................... |
| V | 851 hCoV-19/Japan/PG-2054/2020\|EPI_ISL_685659\|2020-04 | .................... | .................... | ..................... | .................... | .................... | .................... |
| V | 853 hCoV-19/Japan/PG-2056/2020\|EPI_ISL_685661\|2020-04 | .................... | .................... | ..................... | .................... | .................... | .................... |
| V | 854 hCoV-19/Japan/PG-2057/2020\|EPI_ISL_685662\|2020-04 | .................... | .................... | ..................... | .................... | .................... | .................... |
| V | 855 hCoV-19/Japan/PG-2058/2020\|EPI_ISL_685663\|2020-04 | .................... | .................... | ..................... | .................... | .................... | .................... |
| V | 856 hCoV-19/Japan/PG-2059/2020\|EPI_ISL_685664\|2020-04 | .................... | .................... | ..................... | .................... | .................... | .................... |
| V | 857 hCoV-19/Japan/PG-2060/2020\|EPI_ISL_685665\|2020-04 | .................... | .................... | ..................... | .................... | .................... | .................... |
| V | 858 hCoV-19/Japan/PG-2061/2020\|EPI_ISL_685666\|2020-04 | .................... | .................... | ..................... | .................... | .................... | .................... |
| V | 861 hCoV-19/Japan/PG-2064/2020\|EPI_ISL_685669\|2020-04 | .................... | .................... | ..................... | .................... | .................... | .................... |
| V | 862 hCoV-19/Japan/PG-2065/2020\|EPI_ISL_685670\|2020-04 | .................... | .................... | ..................... | .................... | .................... | .................... |
| V | 864 hCoV-19/Japan/PG-2067/2020\|EPI_ISL_685672\|2020-04 | .................... | .................... | ..................... | .................... | .................... | .................... |
| V | 865 hCoV-19/Japan/PG-2068/2020\|EPI_ISL_685673\|2020-04 | .................... | .................... | ..................... | .................... | .................... | .................... |
| V | 866 hCoV-19/Japan/PG-2069/2020\|EPI_ISL_685674\|2020-04 | .................... | .................... | ..................... | .................... | .................... | .................... |
| V | 867 hCoV-19/Japan/PG-2070/2020\|EPI_ISL_685675\|2020-04 | .................... | .................... | ..................... | .................... | .................... | .................... |
| V | 868 hCoV-19/Japan/PG-2071/2020\|EPI_ISL_685676\|2020-04 | .................... | .................... | ..................... | .................... | .................... | .................... |
| V | 869 hCoV-19/Japan/PG-2072/2020\|EPI_ISL_685677\|2020-04 | .................... | .................... | ..................... | .................... | .................... | .................... |
| V | 870 hCoV-19/Japan/PG-2073/2020\|EPI_ISL_685678\|2020-04 | .................... | .................... | ..................... | .................... | .................... | .................... |
| V | 871 hCoV-19/Japan/PG-2074/2020\|EPI_ISL_685679\|2020-04 | .................... | .................... | ..................... | .................... | .................... | .................... |
| V | 872 hCoV-19/Japan/PG-2075/2020\|EPI_ISL_685680\|2020-04 | .................... | .................... | ..................... | .................... | .................... | .................... |
| V | 874 hCoV-19/Japan/PG-2077/2020\|EPI_ISL_685682\|2020-04 | .................... | .................... | ..................... | .................... | .................... | .................... |
| V | 877 hCoV-19/Japan/PG-2080/2020\|EPI_ISL_685685\|2020-04 | .................... | .................... | ..................... | .................... | .................... | .................... |
| V | 878 hCoV-19/Japan/PG-2081/2020\|EPI_ISL_685686\|2020-04 | .................... | .................... | ..................... | .................... | .................... | .................... |
| V | 879 hCoV-19/Japan/PG-2082/2020\|EPI_ISL_685687\|2020-04 | .................... | .................... | ..................... | .................... | .................... | .................... |
| V | 880 hCoV-19/Japan/PG-2083/2020\|EPI_ISL_685688\|2020-04 | .................... | .................... | ..................... | .................... | .................... | .................... |
| V | 881 hCoV-19/Japan/PG-2084/2020\|EPI_ISL_685689\|2020-04 | .................... | .................... | ..................... | .................... | .................... | .................... |
| V | 882 hCoV-19/Japan/PG-2085/2020\|EPI_ISL_685690\|2020-04 | .................... | .................... | ..................... | .................... | .................... | .................... |
| V | 883 hCoV-19/Japan/PG-2086/2020\|EPI_ISL_685691\|2020-04 | .................... | .................... | ..................... | .................... | .................... | .................... |
| V | 884 hCoV-19/Japan/PG-2087/2020\|EPI_ISL_685692\|2020-04 | .................... | .................... | ..................... | .................... | .................... | .................... |
| V | 885 hCoV-19/Japan/PG-2088/2020\|EPI_ISL_685693\|2020-04 | .................... | .................... | ..................... | .................... | .................... | .................... |
| V | 886 hCoV-19/Japan/PG-2089/2020\|EPI_ISL_685694\|2020-04 | .................... | .................... | ..................... | .................... | .................... | .................... |
| V | 887 hCoV-19/Japan/PG-2090/2020\|EPI_ISL_685695\|2020-03 | .................... | .................... | ..................... | .................... | .................... | .................... |
| V | 888 hCoV-19/Japan/PG-2091/2020\|EPI_ISL_685696\|2020-03 | .................... | .................... | ..................... | .................... | .................... | .................... |
| V | 889 hCoV-19/Japan/PG-2092/2020\|EPI_ISL_685697\|2020-03 | .................... | .................... | ..................... | .................... | .................... | .................... |
| V | 890 hCoV-19/Japan/PG-2093/2020\|EPI_ISL_685698\|2020-04 | .................... | .................... | ..................... | .................... | .................... | .................... |
| V | 891 hCoV-19/Japan/PG-2094/2020\|EPI_ISL_685699\|2020-04 | .................... | .................... | ..................... | .................... | .................... | .................... |
| V | 892 hCoV-19/Japan/PG-7873/2020\|EPI_ISL_686974\|2020-07 | .................... | .................... | ..................... | .................... | .................... | .................... |
| V | 893 hCoV-19/Japan/PG-7874/2020\|EPI_ISL_686975\|2020-07 | .................... | .................... | ..................... | .................... | .................... | .................... |
| V | 894 hCoV-19/Japan/PG-7875/2020\|EPI_ISL_686976\|2020-07 | .................... | .................... | ..................... | .................... | .................... | .................... |
| V | 895 hCoV-19/Japan/PG-7876/2020\|EPI_ISL_686977\|2020-07 | .................... | .................... | ..................... | .................... | .................... | .................... |
| V | 896 hCoV-19/Japan/PG-7877/2020\|EPI_ISL_686978\|2020-07 | .................... | .................... | ..................... | .................... | .................... | .................... |
| V | 897 hCoV-19/Japan/PG-7878/2020\|EPI_ISL_686979\|2020-07 | .................... | .................... | ..................... | .................... | .................... | .................... |
| V | 898 hCoV-19/Japan/PG-7879/2020\|EPI_ISL_686980\|2020-07 | .................... | .................... | ..................... | .................... | .................... | .................... |
| V | 899 hCoV-19/Japan/PG-7880/2020\|EPI_ISL_686981\|2020-07 | .................... | .................... | ..................... | .................... | .................... | .................... |
| V | 900 hCoV-19/Japan/PG-7881/2020\|EPI_ISL_686982\|2020-07 | .................... | .................... | ..................... | .................... | .................... | .................... |
| V | 901 hCoV-19/Japan/PG-7882/2020\|EPI_ISL_686983\|2020-07 | .................... | .................... | ..................... | .................... | .................... | .................... |
| V | 902 hCoV-19/Japan/PG-7883/2020\|EPI_ISL_686984\|2020-07 | .................... | .................... | ..................... | .................... | .................... | .................... |
| V | 903 hCoV-19/Japan/PG-7884/2020\|EPI_ISL_686985\|2020-07 | .................... | .................... | ..................... | .................... | .................... | .................... |
| V | 904 hCoV-19/Japan/PG-7885/2020\|EPI_ISL_686986\|2020-07 | .................... | .................... | ..................... | .................... | .................... | .................... |
| V | 905 hCoV-19/Japan/PG-7886/2020\|EPI_ISL_686987\|2020-07 | .................... | .................... | ..................... | .................... | .................... | .................... |
| V | 906 hCoV-19/Japan/PG-7887/2020\|EPI_ISL_686988\|2020-07 | .................... | .................... | ..................... | .................... | .................... | .................... |
| V | 907 hCoV-19/Japan/PG-7888/2020\|EPI_ISL_686989\|2020-07 | .................... | .................... | ..................... | .................... | .................... | .................... |
| V | 908 hCoV-19/Japan/PG-7889/2020\|EPI_ISL_686990\|2020-07 | .................... | .................... | ..................... | .................... | .................... | .................... |
| V | 909 hCoV-19/Japan/PG-7890/2020\|EPI_ISL_686991\|2020-07 | .................... | .................... | ..................... | .................... | .................... | .................... |
| V | 910 hCoV-19/Japan/PG-7891/2020\|EPI_ISL_686992\|2020-07 | .................... | .................... | ..................... | .................... | .................... | .................... |
| V | 911 hCoV-19/Japan/PG-7892/2020\|EPI_ISL_686993\|2020-07 | .................... | .................... | ..................... | .................... | .................... | .................... |
| V | 912 hCoV-19/Japan/PG-7893/2020\|EPI_ISL_686994\|2020-07 | .................... | .................... | ..................... | .................... | .................... | .................... |
| V | 913 hCoV-19/Japan/PG-7894/2020\|EPI_ISL_686995\|2020-07 | .................... | .................... | ..................... | .................... | .................... | .................... |
| V | 914 hCoV-19/Japan/PG-7895/2020\|EPI_ISL_686996\|2020-07 | .................... | .................... | ..................... | .................... | .................... | .................... |
| V | 915 hCoV-19/Japan/PG-7896/2020\|EPI_ISL_686997\|2020-07 | .................... | .................... | ..................... | .................... | .................... | .................... |
| V | 916 hCoV-19/Japan/PG-7897/2020\|EPI_ISL_686998\|2020-07 | .................... | .................... | ..................... | .................... | .................... | .................... |
| V | 917 hCoV-19/Japan/PG-7898/2020\|EPI_ISL_686999\|2020-07 | .................... | .................... | ..................... | .................... | .................... | .................... |
| V | 918 hCoV-19/Japan/PG-4987/2020\|EPI_ISL_690100\|2020-08 | .................... | .................... | ..................... | .................... | .................... | .................... |
| V | 919 hCoV-19/Japan/PG-4988/2020\|EPI_ISL_690101\|2020-08 | .................... | .................... | ..................... | .................... | .................... | .................... |
| V | 920 hCoV-19/Japan/PG-4990/2020\|EPI_ISL_690103\|2020-08 | .................... | .................... | ..................... | .................... | .................... | .................... |
| V | 921 hCoV-19/Japan/PG-4993/2020\|EPI_ISL_690106\|2020-08 | .................... | .................... | ..................... | .................... | .................... | .................... |
| V | 922 hCoV-19/Japan/PG-4994/2020\|EPI_ISL_690107\|2020-08 | .................... | .................... | ..................... | .................... | .................... | .................... |

| | | | | | | | | |
|---|---|---|---|---|---|---|---|---|
| V | 923 | hCoV-19/Japan/PG-4995/2020\|EPI_ISL_690108\|2020-08 | .................... | .................... | .................... | .................... | .................... | .................... |
| V | 924 | hCoV-19/Japan/PG-4996/2020\|EPI_ISL_690109\|2020-08 | .................... | .................... | .................... | .................... | .................... | .................... |
| V | 925 | hCoV-19/Japan/PG-5065/2020\|EPI_ISL_690175\|2020-08 | .................... | .................... | .................... | .................... | .................... | .................... |
| V | 926 | hCoV-19/Japan/PG-5066/2020\|EPI_ISL_690176\|2020-08 | .................... | .................... | .................... | .................... | .................... | .................... |
| V | 927 | hCoV-19/Japan/PG-5067/2020\|EPI_ISL_690177\|2020-08 | .................... | .................... | .................... | .................... | .................... | .................... |
| V | 928 | hCoV-19/Japan/PG-5068/2020\|EPI_ISL_690178\|2020-08 | .................... | .................... | .................... | .................... | .................... | .................... |
| V | 929 | hCoV-19/Japan/PG-5069/2020\|EPI_ISL_690179\|2020-08 | .................... | .................... | .................... | .................... | .................... | .................... |
| V | 930 | hCoV-19/Japan/PG-5070/2020\|EPI_ISL_690180\|2020-08 | .................... | .................... | .................... | .................... | .................... | .................... |
| V | 931 | hCoV-19/Japan/PG-5071/2020\|EPI_ISL_690181\|2020-08 | .................... | .................... | .................... | .................... | .................... | .................... |
| V | 932 | hCoV-19/Japan/PG-5072/2020\|EPI_ISL_690182\|2020-08 | .................... | .................... | .................... | .................... | .................... | .................... |
| V | 933 | hCoV-19/Japan/PG-5073/2020\|EPI_ISL_690183\|2020-08 | .................... | .................... | .................... | .................... | .................... | .................... |
| V | 934 | hCoV-19/Japan/PG-5074/2020\|EPI_ISL_690184\|2020-08 | .................... | .................... | .................... | .................... | .................... | .................... |
| V | 935 | hCoV-19/Japan/PG-5075/2020\|EPI_ISL_690185\|2020-08 | .................... | .................... | .................... | .................... | .................... | .................... |
| V | 936 | hCoV-19/Japan/PG-5076/2020\|EPI_ISL_690186\|2020-08 | .................... | .................... | .................... | .................... | .................... | .................... |
| V | 937 | hCoV-19/Japan/PG-5077/2020\|EPI_ISL_690187\|2020-08 | .................... | .................... | .................... | .................... | .................... | .................... |
| V | 938 | hCoV-19/Japan/PG-5078/2020\|EPI_ISL_690188\|2020-08 | .................... | .................... | .................... | .................... | .................... | .................... |
| V | 939 | hCoV-19/Japan/PG-5079/2020\|EPI_ISL_690189\|2020-08 | .................... | .................... | .................... | .................... | .................... | .................... |
| V | 940 | hCoV-19/Japan/PG-5080/2020\|EPI_ISL_690190\|2020-08 | .................... | .................... | .................... | .................... | .................... | .................... |
| V | 941 | hCoV-19/Japan/PG-5081/2020\|EPI_ISL_690191\|2020-08 | .................... | .................... | .................... | .................... | .................... | .................... |
| V | 942 | hCoV-19/Japan/PG-5082/2020\|EPI_ISL_690192\|2020-08 | .................... | .................... | .................... | .................... | .................... | .................... |
| V | 943 | hCoV-19/Japan/PG-5083/2020\|EPI_ISL_690193\|2020-08 | .................... | .................... | .................... | .................... | .................... | .................... |
| V | 944 | hCoV-19/Japan/PG-5084/2020\|EPI_ISL_690194\|2020-08 | .................... | .................... | .................... | .................... | .................... | .................... |
| V | 945 | hCoV-19/Japan/PG-5085/2020\|EPI_ISL_690195\|2020-08 | .................... | .................... | .................... | .................... | .................... | .................... |
| V | 946 | hCoV-19/Japan/PG-5086/2020\|EPI_ISL_690196\|2020-08 | .................... | .................... | .................... | .................... | .................... | .................... |
| V | 947 | hCoV-19/Japan/PG-5087/2020\|EPI_ISL_690197\|2020-08 | .................... | .................... | .................... | .................... | .................... | .................... |
| V | 948 | hCoV-19/Japan/PG-5088/2020\|EPI_ISL_690198\|2020-08 | .................... | .................... | .................... | .................... | .................... | .................... |
| V | 949 | hCoV-19/Japan/PG-5089/2020\|EPI_ISL_690199\|2020-08 | .................... | .................... | .................... | .................... | .................... | .................... |
| V | 950 | hCoV-19/Japan/PG-3285/2020\|EPI_ISL_691400\|2020-04 | .................... | .................... | .................... | .................... | .................... | .................... |
| V | 951 | hCoV-19/Japan/PG-3286/2020\|EPI_ISL_691401\|2020-04 | .................... | .................... | .................... | .................... | .................... | .................... |
| V | 952 | hCoV-19/Japan/PG-3287/2020\|EPI_ISL_691402\|2020-04 | .................... | .................... | .................... | .................... | .................... | .................... |
| V | 953 | hCoV-19/Japan/PG-3288/2020\|EPI_ISL_691403\|2020-04 | .................... | .................... | .................... | .................... | .................... | .................... |
| V | 954 | hCoV-19/Japan/PG-3289/2020\|EPI_ISL_691404\|2020-04 | .................... | .................... | .................... | .................... | .................... | .................... |
| V | 955 | hCoV-19/Japan/PG-3290/2020\|EPI_ISL_691405\|2020-04 | .................... | .................... | .................... | .................... | .................... | .................... |
| V | 956 | hCoV-19/Japan/PG-3291/2020\|EPI_ISL_691406\|2020-04 | .................... | .................... | .................... | .................... | .................... | .................... |
| V | 957 | hCoV-19/Japan/PG-3292/2020\|EPI_ISL_691407\|2020-04 | .................... | .................... | .................... | .................... | .................... | .................... |
| V | 958 | hCoV-19/Japan/PG-3293/2020\|EPI_ISL_691408\|2020-04 | .................... | .................... | .................... | .................... | .................... | .................... |
| V | 959 | hCoV-19/Japan/PG-3294/2020\|EPI_ISL_691409\|2020-04 | .................... | .................... | .................... | .................... | .................... | .................... |
| V | 960 | hCoV-19/Japan/PG-3295/2020\|EPI_ISL_691410\|2020-04 | .................... | .................... | .................... | .................... | .................... | .................... |
| V | 961 | hCoV-19/Japan/PG-3296/2020\|EPI_ISL_691411\|2020-04 | .................... | .................... | .................... | .................... | .................... | .................... |
| V | 962 | hCoV-19/Japan/PG-3297/2020\|EPI_ISL_691412\|2020-04 | .................... | .................... | .................... | .................... | .................... | .................... |
| V | 963 | hCoV-19/Japan/PG-3298/2020\|EPI_ISL_691413\|2020-04 | .................... | .................... | .................... | .................... | .................... | .................... |
| V | 964 | hCoV-19/Japan/PG-3299/2020\|EPI_ISL_691414\|2020-04 | .................... | .................... | .................... | .................... | .................... | .................... |
| V | 965 | hCoV-19/Japan/PG-3300/2020\|EPI_ISL_691415\|2020-04 | .................... | .................... | .................... | .................... | .................... | .................... |
| V | 966 | hCoV-19/Japan/PG-3301/2020\|EPI_ISL_691416\|2020-04 | .................... | .................... | .................... | .................... | .................... | .................... |
| V | 967 | hCoV-19/Japan/PG-3302/2020\|EPI_ISL_691417\|2020-04 | .................... | .................... | .................... | .................... | .................... | .................... |
| V | 968 | hCoV-19/Japan/PG-3303/2020\|EPI_ISL_691418\|2020-04 | .................... | .................... | .................... | .................... | .................... | .................... |
| V | 969 | hCoV-19/Japan/PG-3304/2020\|EPI_ISL_691419\|2020-04 | .................... | .................... | .................... | .................... | .................... | .................... |
| V | 970 | hCoV-19/Japan/PG-3305/2020\|EPI_ISL_691420\|2020-04 | .................... | .................... | .................... | .................... | .................... | .................... |
| V | 971 | hCoV-19/Japan/PG-3306/2020\|EPI_ISL_691421\|2020-04 | .................... | .................... | .................... | .................... | .................... | .................... |
| V | 972 | hCoV-19/Japan/PG-3307/2020\|EPI_ISL_691422\|2020-04 | .................... | .................... | .................... | .................... | .................... | .................... |
| V | 973 | hCoV-19/Japan/PG-3308/2020\|EPI_ISL_691423\|2020-04 | .................... | .................... | .................... | .................... | .................... | .................... |
| V | 974 | hCoV-19/Japan/PG-3309/2020\|EPI_ISL_691424\|2020-04 | .................... | .................... | .................... | .................... | .................... | .................... |
| V | 975 | hCoV-19/Japan/PG-3310/2020\|EPI_ISL_691425\|2020-04 | .................... | .................... | .................... | .................... | .................... | .................... |
| V | 976 | hCoV-19/Japan/PG-3311/2020\|EPI_ISL_691426\|2020-04 | .................... | .................... | .................... | .................... | .................... | .................... |
| V | 977 | hCoV-19/Japan/PG-3312/2020\|EPI_ISL_691427\|2020-04 | .................... | .................... | .................... | .................... | .................... | .................... |
| V | 979 | hCoV-19/Japan/PG-3314/2020\|EPI_ISL_691429\|2020-04 | .................... | .................... | .................... | .................... | .................... | .................... |
| V | 980 | hCoV-19/Japan/PG-3315/2020\|EPI_ISL_691430\|2020-04 | .................... | .................... | .................... | .................... | .................... | .................... |
| V | 982 | hCoV-19/Japan/PG-3317/2020\|EPI_ISL_691432\|2020-04 | .................... | .................... | .................... | .................... | .................... | .................... |
| V | 983 | hCoV-19/Japan/PG-3320/2020\|EPI_ISL_691435\|2020-04 | .................... | .................... | .................... | .................... | .................... | .................... |
| V | 984 | hCoV-19/Japan/PG-3321/2020\|EPI_ISL_691436\|2020-04 | .................... | .................... | .................... | .................... | .................... | .................... |
| V | 985 | hCoV-19/Japan/PG-3322/2020\|EPI_ISL_691437\|2020-04 | .................... | .................... | .................... | .................... | .................... | .................... |
| V | 986 | hCoV-19/Japan/PG-3323/2020\|EPI_ISL_691438\|2020-04 | .................... | .................... | .................... | .................... | .................... | .................... |
| V | 987 | hCoV-19/Japan/PG-3324/2020\|EPI_ISL_691439\|2020-04 | .................... | .................... | .................... | .................... | .................... | .................... |
| V | 988 | hCoV-19/Japan/PG-4507/2020\|EPI_ISL_692730\|2020-07 | .................... | .................... | .................... | .................... | .................... | .................... |
| V | 989 | hCoV-19/Japan/PG-4508/2020\|EPI_ISL_692731\|2020-07 | .................... | .................... | .................... | .................... | .................... | .................... |
| V | 990 | hCoV-19/Japan/PG-4509/2020\|EPI_ISL_692732\|2020-07 | .................... | .................... | .................... | .................... | .................... | .................... |
| V | 991 | hCoV-19/France/ARA-57686/2020\|EPI_ISL_692733\|2020- | .................... | .................... | .................... | .................... | .................... | .................... |
| V | 992 | hCoV-19/France/ARA-59684/2020\|EPI_ISL_692734\|2020- | .................... | .................... | .................... | .................... | .................... | .................... |
| V | 993 | hCoV-19/France/ARA-59685/2020\|EPI_ISL_692735\|2020- | .................... | .................... | .................... | .................... | .................... | .................... |
| V | 994 | hCoV-19/France/ARA-61889/2020\|EPI_ISL_692736\|2020- | .................... | .................... | .................... | .................... | .................... | .................... |
| V | 995 | hCoV-19/France/ARA-66327/2020\|EPI_ISL_692737\|2020- | .................... | .................... | .................... | .................... | .................... | .................... |
| V | 996 | hCoV-19/France/ARA-66446/2020\|EPI_ISL_692738\|2020- | .................... | .................... | .................... | .................... | .................... | .................... |
| V | 997 | hCoV-19/France/ARA-67140/2020\|EPI_ISL_692739\|2020- | .................... | .................... | .................... | .................... | .................... | .................... |
| V | 998 | hCoV-19/France/ARA-67616/2020\|EPI_ISL_692740\|2020- | .................... | .................... | .................... | .................... | .................... | .................... |
| V | 999 | hCoV-19/France/ARA-67659/2020\|EPI_ISL_692741\|2020- | .................... | .................... | .................... | .................... | .................... | .................... |
| V | 1000 | hCoV-19/France/ARA-67716/2020\|EPI_ISL_692742\|2020- | .................... | .................... | .................... | .................... | .................... | .................... |
| V | 1001 | hCoV-19/France/ARA-6773/2020\|EPI_ISL_692743\|2020-0 | .................... | .................... | .................... | .................... | .................... | .................... |
| V | 1002 | hCoV-19/France/ARA-68122/2020\|EPI_ISL_692744\|2020- | .................... | .................... | .................... | .................... | .................... | .................... |
| V | 1003 | hCoV-19/France/ARA-70123/2020\|EPI_ISL_692745\|2020- | .................... | .................... | .................... | .................... | .................... | .................... |
| V | 1004 | hCoV-19/France/ARA-71437/2020\|EPI_ISL_692746\|2020- | .................... | .................... | .................... | .................... | .................... | .................... |

| | | | | | | | | |
|---|---|---|---|---|---|---|---|---|
| V | 1005 | hCoV-19/France/ARA-71440/2020\|EPI_ISL_692747\|2020- | .................... | .................... | .................... | .................... | .................... | .................... |
| V | 1006 | hCoV-19/France/ARA-71476/2020\|EPI_ISL_692748\|2020- | .................... | .................... | .................... | .................... | .................... | .................... |
| V | 1007 | hCoV-19/France/ARA-71532/2020\|EPI_ISL_692749\|2020- | .................... | .................... | .................... | .................... | .................... | .................... |
| V | 1008 | hCoV-19/France/ARA-72111/2020\|EPI_ISL_692750\|2020- | .................... | .................... | .................... | .................... | .................... | .................... |
| V | 1009 | hCoV-19/France/ARA-7212/2020\|EPI_ISL_692751\|2020-0 | .................... | .................... | .................... | .................... | .................... | .................... |
| V | 1010 | hCoV-19/France/ARA-7215/2020\|EPI_ISL_692752\|2020-0 | .................... | .................... | .................... | .................... | .................... | .................... |
| V | 1011 | hCoV-19/France/ARA-73624/2020\|EPI_ISL_692753\|2020- | .................... | .................... | .................... | .................... | .................... | .................... |
| V | 1012 | hCoV-19/France/ARA-74140/2020\|EPI_ISL_692754\|2020- | .................... | .................... | .................... | .................... | .................... | .................... |
| V | 1013 | hCoV-19/France/ARA-57804/2020\|EPI_ISL_692755\|2020- | .................... | .................... | .................... | .................... | .................... | .................... |
| V | 1014 | hCoV-19/France/ARA-62561/2020\|EPI_ISL_692756\|2020- | .................... | .................... | .................... | .................... | .................... | .................... |
| V | 1015 | hCoV-19/France/ARA-62602/2020\|EPI_ISL_692757\|2020- | .................... | .................... | .................... | .................... | .................... | .................... |
| V | 1016 | hCoV-19/France/ARA-62907/2020\|EPI_ISL_692758\|2020- | .................... | .................... | .................... | .................... | .................... | .................... |
| V | 1017 | hCoV-19/France/ARA-63103/2020\|EPI_ISL_692759\|2020- | .................... | .................... | .................... | .................... | .................... | .................... |
| V | 1018 | hCoV-19/France/ARA-64304/2020\|EPI_ISL_692760\|2020- | .................... | .................... | .................... | .................... | .................... | .................... |
| V | 1019 | hCoV-19/France/ARA-65499/2020\|EPI_ISL_692761\|2020- | .................... | .................... | .................... | .................... | .................... | .................... |
| V | 1020 | hCoV-19/France/ARA-66427/2020\|EPI_ISL_692762\|2020- | .................... | .................... | .................... | .................... | .................... | .................... |
| V | 1021 | hCoV-19/France/ARA-6761/2020\|EPI_ISL_692763\|2020-0 | .................... | .................... | .................... | .................... | .................... | .................... |
| V | 1022 | hCoV-19/France/ARA-67717/2020\|EPI_ISL_692766\|2020- | .................... | .................... | .................... | .................... | .................... | .................... |
| V | 1023 | hCoV-19/France/ARA-68373/2020\|EPI_ISL_692768\|2020- | .................... | .................... | .................... | .................... | .................... | .................... |
| V | 1024 | hCoV-19/USA/MA-MASPHL-01093/2020\|EPI_ISL_692769\|20 | .................... | .................... | .................... | .................... | .................... | .................... |
| V | 1025 | hCoV-19/Japan/PG-0669/2020\|EPI_ISL_684279\|2020-04 | .................... | .................... | .................... | .................... | .................... | .................... |
| V | 1026 | hCoV-19/Japan/PG-0670/2020\|EPI_ISL_684280\|2020-04 | .................... | .................... | .................... | .................... | .................... | .................... |
| V | 1027 | hCoV-19/Japan/PG-0671/2020\|EPI_ISL_684281\|2020-04 | .................... | .................... | .................... | .................... | .................... | .................... |
| V | 1028 | hCoV-19/Japan/PG-0672/2020\|EPI_ISL_684282\|2020-04 | .................... | .................... | .................... | .................... | .................... | .................... |
| V | 1029 | hCoV-19/Japan/PG-0673/2020\|EPI_ISL_684283\|2020-04 | .................... | .................... | .................... | .................... | .................... | .................... |
| V | 1030 | hCoV-19/Japan/PG-0674/2020\|EPI_ISL_684284\|2020-04 | .................... | .................... | .................... | .................... | .................... | .................... |
| V | 1031 | hCoV-19/Japan/PG-0675/2020\|EPI_ISL_684285\|2020-04 | .................... | .................... | .................... | .................... | .................... | .................... |
| V | 1032 | hCoV-19/Japan/PG-0676/2020\|EPI_ISL_684286\|2020-04 | .................... | .................... | .................... | .................... | .................... | .................... |
| V | 1033 | hCoV-19/Japan/PG-0677/2020\|EPI_ISL_684287\|2020-04 | .................... | .................... | .................... | .................... | .................... | .................... |
| V | 1034 | hCoV-19/Japan/PG-0678/2020\|EPI_ISL_684288\|2020-04 | .................... | .................... | .................... | .................... | .................... | .................... |
| V | 1035 | hCoV-19/Japan/PG-0679/2020\|EPI_ISL_684289\|2020-04 | .................... | .................... | .................... | .................... | .................... | .................... |
| V | 1036 | hCoV-19/Japan/PG-0680/2020\|EPI_ISL_684290\|2020-04 | .................... | .................... | .................... | .................... | .................... | .................... |
| V | 1037 | hCoV-19/Japan/PG-0681/2020\|EPI_ISL_684291\|2020-04 | .................... | .................... | .................... | .................... | .................... | .................... |
| V | 1038 | hCoV-19/Japan/PG-0682/2020\|EPI_ISL_684292\|2020-04 | .................... | .................... | .................... | .................... | .................... | .................... |
| V | 1039 | hCoV-19/Japan/PG-0683/2020\|EPI_ISL_684293\|2020-04 | .................... | .................... | .................... | .................... | .................... | .................... |
| V | 1040 | hCoV-19/Japan/PG-0684/2020\|EPI_ISL_684294\|2020-04 | .................... | .................... | .................... | .................... | .................... | .................... |
| V | 1041 | hCoV-19/Japan/PG-0685/2020\|EPI_ISL_684295\|2020-04 | .................... | .................... | .................... | .................... | .................... | .................... |
| V | 1042 | hCoV-19/Japan/PG-0686/2020\|EPI_ISL_684296\|2020 | .................... | .................... | .................... | .................... | .................... | .................... |
| V | 1043 | hCoV-19/Japan/PG-0687/2020\|EPI_ISL_684297\|2020-04 | .................... | .................... | .................... | .................... | .................... | .................... |
| V | 1044 | hCoV-19/Japan/PG-0688/2020\|EPI_ISL_684298\|2020-04 | .................... | .................... | .................... | .................... | .................... | .................... |
| V | 1045 | hCoV-19/Japan/PG-0689/2020\|EPI_ISL_684299\|2020-04 | .................... | .................... | .................... | .................... | .................... | .................... |
| V | 1046 | hCoV-19/Japan/PG-1905/2020\|EPI_ISL_685511\|2020-04 | .................... | .................... | .................... | .................... | .................... | .................... |
| V | 1047 | hCoV-19/Japan/PG-1906/2020\|EPI_ISL_685512\|2020-04 | .................... | .................... | .................... | .................... | .................... | .................... |
| V | 1048 | hCoV-19/Japan/PG-1907/2020\|EPI_ISL_685513\|2020-04 | .................... | .................... | .................... | .................... | .................... | .................... |
| V | 1050 | hCoV-19/Japan/PG-1909/2020\|EPI_ISL_685515\|2020-04 | .................... | .................... | .................... | .................... | .................... | .................... |
| V | 1051 | hCoV-19/Japan/PG-1910/2020\|EPI_ISL_685516\|2020-04 | .................... | .................... | .................... | .................... | .................... | .................... |
| V | 1052 | hCoV-19/Japan/PG-1911/2020\|EPI_ISL_685517\|2020-04 | .................... | .................... | .................... | .................... | .................... | .................... |
| V | 1053 | hCoV-19/Japan/PG-1912/2020\|EPI_ISL_685518\|2020-04 | .................... | .................... | .................... | .................... | .................... | .................... |
| V | 1054 | hCoV-19/Japan/PG-1913/2020\|EPI_ISL_685519\|2020-04 | .................... | .................... | .................... | .................... | .................... | .................... |
| V | 1055 | hCoV-19/Japan/PG-1914/2020\|EPI_ISL_685520\|2020-04 | .................... | .................... | .................... | .................... | .................... | .................... |
| V | 1056 | hCoV-19/Japan/PG-1915/2020\|EPI_ISL_685521\|2020-05 | .................... | .................... | .................... | .................... | .................... | .................... |
| V | 1057 | hCoV-19/Japan/PG-7738/2020\|EPI_ISL_686840\|2020-09 | .................... | .................... | .................... | .................... | .................... | .................... |
| V | 1058 | hCoV-19/Japan/PG-7740/2020\|EPI_ISL_686842\|2020-09 | .................... | .................... | .................... | .................... | .................... | .................... |
| V | 1059 | hCoV-19/Japan/PG-7741/2020\|EPI_ISL_686843\|2020-09 | .................... | .................... | .................... | .................... | .................... | .................... |
| V | 1060 | hCoV-19/Japan/PG-7742/2020\|EPI_ISL_686844\|2020-09 | .................... | .................... | .................... | .................... | .................... | .................... |
| V | 1061 | hCoV-19/Japan/PG-7743/2020\|EPI_ISL_686845\|2020-03 | .................... | .................... | .................... | .................... | .................... | .................... |
| V | 1062 | hCoV-19/Japan/PG-7744/2020\|EPI_ISL_686846\|2020-03 | .................... | .................... | .................... | .................... | .................... | .................... |
| V | 1063 | hCoV-19/Japan/PG-7745/2020\|EPI_ISL_686847\|2020-03 | .................... | .................... | .................... | .................... | .................... | .................... |
| V | 1064 | hCoV-19/Japan/PG-7746/2020\|EPI_ISL_686848\|2020-03 | .................... | .................... | .................... | .................... | .................... | .................... |
| V | 1065 | hCoV-19/Japan/PG-7747/2020\|EPI_ISL_686849\|2020-04 | .................... | .................... | .................... | .................... | .................... | .................... |
| V | 1066 | hCoV-19/Japan/PG-7748/2020\|EPI_ISL_686850\|2020-04 | .................... | .................... | .................... | .................... | .................... | .................... |
| V | 1067 | hCoV-19/Japan/PG-7749/2020\|EPI_ISL_686851\|2020-04 | .................... | .................... | .................... | .................... | .................... | .................... |
| V | 1068 | hCoV-19/Japan/PG-7750/2020\|EPI_ISL_686852\|2020-04 | .................... | .................... | .................... | .................... | .................... | .................... |
| V | 1069 | hCoV-19/Japan/PG-5987/2020\|EPI_ISL_689100\|2020-08 | .................... | .................... | .................... | .................... | .................... | .................... |
| V | 1070 | hCoV-19/Japan/PG-5988/2020\|EPI_ISL_689101\|2020-08 | .................... | .................... | .................... | .................... | .................... | .................... |
| V | 1071 | hCoV-19/Japan/PG-5989/2020\|EPI_ISL_689102\|2020-08 | .................... | .................... | .................... | .................... | .................... | .................... |
| V | 1072 | hCoV-19/Japan/PG-5990/2020\|EPI_ISL_689103\|2020-08 | .................... | .................... | .................... | .................... | .................... | .................... |
| V | 1073 | hCoV-19/Japan/PG-5991/2020\|EPI_ISL_689104\|2020-07 | .................... | .................... | .................... | .................... | .................... | .................... |
| V | 1074 | hCoV-19/Japan/PG-5992/2020\|EPI_ISL_689105\|2020-07 | .................... | .................... | .................... | .................... | .................... | .................... |
| V | 1075 | hCoV-19/Japan/PG-5993/2020\|EPI_ISL_689106\|2020-07 | .................... | .................... | .................... | .................... | .................... | .................... |
| V | 1076 | hCoV-19/Japan/PG-5994/2020\|EPI_ISL_689107\|2020-07 | .................... | .................... | .................... | .................... | .................... | .................... |
| V | 1077 | hCoV-19/Japan/PG-5995/2020\|EPI_ISL_689108\|2020-07 | .................... | .................... | .................... | .................... | .................... | .................... |
| V | 1078 | hCoV-19/Japan/PG-5996/2020\|EPI_ISL_689109\|2020-07 | .................... | .................... | .................... | .................... | .................... | .................... |
| V | 1079 | hCoV-19/Japan/PG-5997/2020\|EPI_ISL_689110\|2020-07 | .................... | .................... | .................... | .................... | .................... | .................... |
| V | 1080 | hCoV-19/Japan/PG-5998/2020\|EPI_ISL_689111\|2020-07 | .................... | .................... | .................... | .................... | .................... | .................... |
| V | 1081 | hCoV-19/Japan/PG-5999/2020\|EPI_ISL_689112\|2020-07 | .................... | .................... | .................... | .................... | .................... | .................... |
| V | 1082 | hCoV-19/Japan/PG-6000/2020\|EPI_ISL_689113\|2020-07 | .................... | .................... | .................... | .................... | .................... | .................... |
| V | 1083 | hCoV-19/Japan/PG-6001/2020\|EPI_ISL_689114\|2020-07 | .................... | .................... | .................... | .................... | .................... | .................... |
| V | 1084 | hCoV-19/Japan/PG-6002/2020\|EPI_ISL_689115\|2020-07 | .................... | .................... | .................... | .................... | .................... | .................... |
| V | 1085 | hCoV-19/Japan/PG-6003/2020\|EPI_ISL_689116\|2020-07 | .................... | .................... | .................... | .................... | .................... | .................... |

V 1086 hCoV-19/Japan/PG-6004/2020|EPI_ISL_689117|2020-07 .................... .................... ..................... .................... .................... ....................
V 1087 hCoV-19/Japan/PG-6005/2020|EPI_ISL_689118|2020-07 .................... .................... ..................... .................... .................... ....................
V 1088 hCoV-19/Japan/PG-6006/2020|EPI_ISL_689119|2020-07 .................... .................... ..................... .................... .................... ....................
V 1089 hCoV-19/Japan/PG-6007/2020|EPI_ISL_689120|2020-07 .................... .................... ..................... .................... .................... ....................
V 1090 hCoV-19/Japan/PG-6008/2020|EPI_ISL_689121|2020-07 .................... .................... ..................... .................... .................... ....................
V 1091 hCoV-19/Japan/PG-6009/2020|EPI_ISL_689122|2020-07 .................... .................... ..................... .................... .................... ....................
V 1092 hCoV-19/Japan/PG-6010/2020|EPI_ISL_689123|2020-07 .................... .................... ..................... .................... .................... ....................
V 1093 hCoV-19/Japan/PG-6011/2020|EPI_ISL_689124|2020-07 .................... .................... ..................... .................... .................... ....................
V 1094 hCoV-19/Japan/PG-6012/2020|EPI_ISL_689125|2020-07 .................... .................... ..................... .................... .................... ....................
V 1095 hCoV-19/Japan/PG-6013/2020|EPI_ISL_689126|2020-07 .................... .................... ..................... .................... .................... ....................
V 1096 hCoV-19/Japan/PG-6014/2020|EPI_ISL_689127|2020-07 .................... .................... ..................... .................... .................... ....................
V 1097 hCoV-19/Japan/PG-6016/2020|EPI_ISL_689128|2020-07 .................... .................... ..................... .................... .................... ....................
V 1098 hCoV-19/Japan/PG-6017/2020|EPI_ISL_689129|2020-07 .................... .................... ..................... .................... .................... ....................
V 1099 hCoV-19/Japan/PG-6018/2020|EPI_ISL_689130|2020-07 .................... .................... ..................... .................... .................... ....................
V 1100 hCoV-19/Japan/PG-6019/2020|EPI_ISL_689131|2020-07 .................... .................... ..................... .................... .................... ....................
V 1101 hCoV-19/Japan/PG-6020/2020|EPI_ISL_689132|2020-07 .................... .................... ..................... .................... .................... ....................
V 1102 hCoV-19/Japan/PG-6021/2020|EPI_ISL_689133|2020-07 .................... .................... ..................... .................... .................... ....................
V 1103 hCoV-19/Japan/PG-6022/2020|EPI_ISL_689134|2020-07 .................... .................... ..................... .................... .................... ....................
V 1104 hCoV-19/Japan/PG-6023/2020|EPI_ISL_689135|2020-07 .................... .................... ..................... .................... .................... ....................
V 1105 hCoV-19/Japan/PG-6024/2020|EPI_ISL_689136|2020-07 .................... .................... ..................... .................... .................... ....................
V 1106 hCoV-19/Japan/PG-6025/2020|EPI_ISL_689137|2020-07 .................... .................... ..................... .................... .................... ....................
V 1107 hCoV-19/Japan/PG-6026/2020|EPI_ISL_689138|2020-07 .................... .................... ..................... .................... .................... ....................
V 1108 hCoV-19/Japan/PG-6027/2020|EPI_ISL_689139|2020-07 .................... .................... ..................... .................... .................... ....................
V 1109 hCoV-19/Japan/PG-6028/2020|EPI_ISL_689140|2020-07 .................... .................... ..................... .................... .................... ....................
V 1110 hCoV-19/Japan/PG-6029/2020|EPI_ISL_689141|2020-07 .................... .................... ..................... .................... .................... ....................
V 1111 hCoV-19/Japan/PG-6030/2020|EPI_ISL_689142|2020-07 .................... .................... ..................... .................... .................... ....................
V 1112 hCoV-19/Japan/PG-6031/2020|EPI_ISL_689143|2020-07 .................... .................... ..................... .................... .................... ....................
V 1113 hCoV-19/Japan/PG-6032/2020|EPI_ISL_689144|2020-07 .................... .................... ..................... .................... .................... ....................
V 1114 hCoV-19/Japan/PG-6033/2020|EPI_ISL_689145|2020-07 .................... .................... ..................... .................... .................... ....................
V 1115 hCoV-19/Japan/PG-6034/2020|EPI_ISL_689146|2020-07 .................... .................... ..................... .................... .................... ....................
V 1116 hCoV-19/Japan/PG-6035/2020|EPI_ISL_689147|2020-07 .................... .................... ..................... .................... .................... ....................
V 1117 hCoV-19/Japan/PG-6036/2020|EPI_ISL_689148|2020-07 .................... .................... ..................... .................... .................... ....................
V 1118 hCoV-19/Japan/PG-6037/2020|EPI_ISL_689149|2020-07 .................... .................... ..................... .................... .................... ....................
V 1119 hCoV-19/Japan/PG-6038/2020|EPI_ISL_689150|2020-07 .................... .................... ..................... .................... .................... ....................
V 1120 hCoV-19/Japan/PG-6039/2020|EPI_ISL_689151|2020-07 .................... .................... ..................... .................... .................... ....................
V 1121 hCoV-19/Japan/PG-6040/2020|EPI_ISL_689152|2020-07 .................... .................... ..................... .................... .................... ....................
V 1122 hCoV-19/Japan/PG-6041/2020|EPI_ISL_689153|2020-07 .................... .................... ..................... .................... .................... ....................
V 1123 hCoV-19/Japan/PG-6042/2020|EPI_ISL_689154|2020-07 .................... .................... ..................... .................... .................... ....................
V 1124 hCoV-19/Japan/PG-6043/2020|EPI_ISL_689155|2020-07 .................... .................... ..................... .................... .................... ....................
V 1125 hCoV-19/Japan/PG-6044/2020|EPI_ISL_689156|2020-07 .................... .................... ..................... .................... .................... ....................
V 1126 hCoV-19/Japan/PG-6045/2020|EPI_ISL_689157|2020-07 .................... .................... ..................... .................... .................... ....................
V 1127 hCoV-19/Japan/PG-6046/2020|EPI_ISL_689158|2020-07 .................... .................... ..................... .................... .................... ....................
V 1128 hCoV-19/Japan/PG-6047/2020|EPI_ISL_689159|2020-07 .................... .................... ..................... .................... .................... ....................
V 1129 hCoV-19/Japan/PG-6048/2020|EPI_ISL_689160|2020-07 .................... .................... ..................... .................... .................... ....................
V 1130 hCoV-19/Japan/PG-6049/2020|EPI_ISL_689161|2020-07 .................... .................... ..................... .................... .................... ....................
V 1131 hCoV-19/Japan/PG-6050/2020|EPI_ISL_689162|2020-07 .................... .................... ..................... .................... .................... ....................
V 1132 hCoV-19/Japan/PG-6051/2020|EPI_ISL_689163|2020-07 .................... .................... ..................... .................... .................... ....................
V 1133 hCoV-19/Japan/PG-6052/2020|EPI_ISL_689164|2020-07 .................... .................... ..................... .................... .................... ....................
V 1134 hCoV-19/Japan/PG-6053/2020|EPI_ISL_689165|2020-07 .................... .................... ..................... .................... .................... ....................
V 1135 hCoV-19/Japan/PG-6054/2020|EPI_ISL_689166|2020-07 .................... .................... ..................... .................... .................... ....................
V 1136 hCoV-19/Japan/PG-6055/2020|EPI_ISL_689167|2020-07 .................... .................... ..................... .................... .................... ....................
V 1137 hCoV-19/Japan/PG-6056/2020|EPI_ISL_689168|2020-07 .................... .................... ..................... .................... .................... ....................
V 1138 hCoV-19/Japan/PG-6057/2020|EPI_ISL_689169|2020-07 .................... .................... ..................... .................... .................... ....................
V 1139 hCoV-19/Japan/PG-4989/2020|EPI_ISL_690102|2020-08 .................... .................... ..................... .................... .................... ....................
V 1140 hCoV-19/Japan/PG-4991/2020|EPI_ISL_690104|2020-08 .................... .................... ..................... .................... .................... ....................
V 1141 hCoV-19/Japan/PG-4992/2020|EPI_ISL_690105|2020-08 .................... .................... ..................... .................... .................... ....................
V 1142 hCoV-19/Japan/PG-4997/2020|EPI_ISL_690110|2020-08 .................... .................... ..................... .................... .................... ....................
V 1143 hCoV-19/Japan/PG-4998/2020|EPI_ISL_690111|2020-08 .................... .................... ..................... .................... .................... ....................
V 1144 hCoV-19/Japan/PG-4999/2020|EPI_ISL_690112|2020-08 .................... .................... ..................... .................... .................... ....................
V 1145 hCoV-19/Japan/PG-5000/2020|EPI_ISL_690113|2020-08 .................... .................... ..................... .................... .................... ....................
V 1146 hCoV-19/Japan/PG-5001/2020|EPI_ISL_690114|2020-08 .................... .................... ..................... .................... .................... ....................
V 1147 hCoV-19/Japan/PG-5002/2020|EPI_ISL_690115|2020-08 .................... .................... ..................... .................... .................... ....................
V 1148 hCoV-19/Japan/PG-5003/2020|EPI_ISL_690116|2020-08 .................... .................... ..................... .................... .................... ....................
V 1149 hCoV-19/Japan/PG-5004/2020|EPI_ISL_690117|2020-08 .................... .................... ..................... .................... .................... ....................
V 1150 hCoV-19/Japan/PG-5005/2020|EPI_ISL_690118|2020-08 .................... .................... ..................... .................... .................... ....................
V 1151 hCoV-19/Japan/PG-5006/2020|EPI_ISL_690119|2020-08 .................... .................... ..................... .................... .................... ....................
V 1152 hCoV-19/Japan/PG-5008/2020|EPI_ISL_690120|2020-08 .................... .................... ..................... .................... .................... ....................
V 1153 hCoV-19/Japan/PG-5009/2020|EPI_ISL_690121|2020-08 .................... .................... ..................... .................... .................... ....................
V 1154 hCoV-19/Japan/PG-5010/2020|EPI_ISL_690122|2020-08 .................... .................... ..................... .................... .................... ....................
V 1155 hCoV-19/Japan/PG-5012/2020|EPI_ISL_690123|2020-08 .................... .................... ..................... .................... .................... ....................
V 1156 hCoV-19/Japan/PG-5013/2020|EPI_ISL_690124|2020-08 .................... .................... ..................... .................... .................... ....................
V 1157 hCoV-19/Japan/PG-5014/2020|EPI_ISL_690125|2020-08 .................... .................... ..................... .................... .................... ....................
V 1158 hCoV-19/Japan/PG-5015/2020|EPI_ISL_690126|2020-08 .................... .................... ..................... .................... .................... ....................
V 1159 hCoV-19/Japan/PG-5016/2020|EPI_ISL_690127|2020-08 .................... .................... ..................... .................... .................... ....................
V 1160 hCoV-19/Japan/PG-5017/2020|EPI_ISL_690128|2020-08 .................... .................... ..................... .................... .................... ....................
V 1161 hCoV-19/Japan/PG-5018/2020|EPI_ISL_690129|2020-08 .................... .................... ..................... .................... .................... ....................
V 1162 hCoV-19/Japan/PG-5019/2020|EPI_ISL_690130|2020-08 .................... .................... ..................... .................... .................... ....................
V 1163 hCoV-19/Japan/PG-5020/2020|EPI_ISL_690131|2020-08 .................... .................... ..................... .................... .................... ....................
V 1164 hCoV-19/Japan/PG-5021/2020|EPI_ISL_690132|2020-08 .................... .................... ..................... .................... .................... ....................
V 1165 hCoV-19/Japan/PG-5022/2020|EPI_ISL_690133|2020-08 .................... .................... ..................... .................... .................... ....................

| | | | | | | | | |
|---|---|---|---|---|---|---|---|---|
| V | 1166 | hCoV-19/Japan/PG-5023/2020\|EPI_ISL_690134\|2020-08 | .................... | .................... | ..................... | .................... | .................... | .................... |
| V | 1167 | hCoV-19/Japan/PG-5024/2020\|EPI_ISL_690135\|2020-08 | .................... | .................... | ..................... | .................... | .................... | .................... |
| V | 1168 | hCoV-19/Japan/PG-5025/2020\|EPI_ISL_690136\|2020-08 | .................... | .................... | ..................... | .................... | .................... | .................... |
| V | 1169 | hCoV-19/Japan/PG-5026/2020\|EPI_ISL_690137\|2020-07 | .................... | .................... | ..................... | .................... | .................... | .................... |
| V | 1170 | hCoV-19/Japan/PG-5027/2020\|EPI_ISL_690138\|2020-07 | .................... | .................... | ..................... | .................... | .................... | .................... |
| V | 1171 | hCoV-19/Japan/PG-5028/2020\|EPI_ISL_690139\|2020-07 | .................... | .................... | ..................... | .................... | .................... | .................... |
| V | 1172 | hCoV-19/Japan/PG-5029/2020\|EPI_ISL_690140\|2020-07 | .................... | .................... | ..................... | .................... | .................... | .................... |
| V | 1173 | hCoV-19/Japan/PG-5030/2020\|EPI_ISL_690141\|2020-07 | .................... | .................... | ..................... | .................... | .................... | .................... |
| V | 1175 | hCoV-19/Japan/PG-5032/2020\|EPI_ISL_690143\|2020-07 | .................... | .................... | ..................... | .................... | .................... | .................... |
| V | 1176 | hCoV-19/Japan/PG-5033/2020\|EPI_ISL_690144\|2020-07 | .................... | .................... | ..................... | .................... | .................... | .................... |
| V | 1177 | hCoV-19/Japan/PG-5034/2020\|EPI_ISL_690145\|2020-07 | .................... | .................... | ..................... | .................... | .................... | .................... |
| V | 1178 | hCoV-19/Japan/PG-5035/2020\|EPI_ISL_690146\|2020-07 | .................... | .................... | ..................... | .................... | .................... | .................... |
| V | 1179 | hCoV-19/Japan/PG-5036/2020\|EPI_ISL_690147\|2020-07 | .................... | .................... | ..................... | .................... | .................... | .................... |
| V | 1180 | hCoV-19/Japan/PG-5037/2020\|EPI_ISL_690148\|2020-07 | .................... | .................... | ..................... | .................... | .................... | .................... |
| V | 1181 | hCoV-19/Japan/PG-5038/2020\|EPI_ISL_690149\|2020-07 | .................... | .................... | ..................... | .................... | .................... | .................... |
| V | 1182 | hCoV-19/Japan/PG-5039/2020\|EPI_ISL_690150\|2020-07 | .................... | .................... | ..................... | .................... | .................... | .................... |
| V | 1183 | hCoV-19/Japan/PG-5040/2020\|EPI_ISL_690151\|2020-07 | .................... | .................... | ..................... | .................... | .................... | .................... |
| V | 1184 | hCoV-19/Japan/PG-5041/2020\|EPI_ISL_690152\|2020-07 | .................... | .................... | ..................... | .................... | .................... | .................... |
| V | 1185 | hCoV-19/Japan/PG-3318/2020\|EPI_ISL_691433\|2020-04 | .................... | .................... | ..................... | .................... | .................... | .................... |
| V | 1186 | hCoV-19/Japan/PG-3319/2020\|EPI_ISL_691434\|2020-04 | .................... | .................... | ..................... | .................... | .................... | .................... |
| V | 1188 | hCoV-19/Japan/PG-3326/2020\|EPI_ISL_691441\|2020-04 | .................... | .................... | ..................... | .................... | .................... | .................... |
| V | 1189 | hCoV-19/Japan/PG-3327/2020\|EPI_ISL_691442\|2020-04 | .................... | .................... | ..................... | .................... | .................... | .................... |
| V | 1190 | hCoV-19/Japan/PG-3328/2020\|EPI_ISL_691443\|2020-04 | .................... | .................... | ..................... | .................... | .................... | .................... |
| V | 1191 | hCoV-19/Japan/PG-3329/2020\|EPI_ISL_691444\|2020-04 | .................... | .................... | ..................... | .................... | .................... | .................... |
| V | 1192 | hCoV-19/Japan/PG-3330/2020\|EPI_ISL_691445\|2020-04 | .................... | .................... | ..................... | .................... | .................... | .................... |
| V | 1193 | hCoV-19/Japan/PG-3331/2020\|EPI_ISL_691446\|2020-04 | .................... | .................... | ..................... | .................... | .................... | .................... |
| V | 1194 | hCoV-19/Japan/PG-3332/2020\|EPI_ISL_691447\|2020-04 | .................... | .................... | ..................... | .................... | .................... | .................... |
| V | 1195 | hCoV-19/Japan/PG-3333/2020\|EPI_ISL_691448\|2020-04 | .................... | .................... | ..................... | .................... | .................... | .................... |
| V | 1196 | hCoV-19/Japan/PG-3334/2020\|EPI_ISL_691449\|2020-04 | .................... | .................... | ..................... | .................... | .................... | .................... |
| V | 1197 | hCoV-19/Japan/PG-3335/2020\|EPI_ISL_691450\|2020-04 | .................... | .................... | ..................... | .................... | .................... | .................... |
| V | 1198 | hCoV-19/Japan/PG-3336/2020\|EPI_ISL_691451\|2020-04 | .................... | .................... | ..................... | .................... | .................... | .................... |
| V | 1199 | hCoV-19/Japan/PG-3337/2020\|EPI_ISL_691452\|2020-04 | .................... | .................... | ..................... | .................... | .................... | .................... |
| V | 1200 | hCoV-19/Japan/PG-3338/2020\|EPI_ISL_691453\|2020-04 | .................... | .................... | ..................... | .................... | .................... | .................... |
| V | 1201 | hCoV-19/Japan/PG-3339/2020\|EPI_ISL_691454\|2020-04 | .................... | .................... | ..................... | .................... | .................... | .................... |
| V | 1202 | hCoV-19/Japan/PG-3340/2020\|EPI_ISL_691455\|2020-04 | .................... | .................... | ..................... | .................... | .................... | .................... |
| V | 1203 | hCoV-19/Japan/PG-3341/2020\|EPI_ISL_691456\|2020-04 | .................... | .................... | ..................... | .................... | .................... | .................... |
| V | 1204 | hCoV-19/Japan/PG-3342/2020\|EPI_ISL_691457\|2020-04 | .................... | .................... | ..................... | .................... | .................... | .................... |
| V | 1205 | hCoV-19/Japan/PG-3343/2020\|EPI_ISL_691458\|2020-04 | .................... | .................... | ..................... | .................... | .................... | .................... |
| V | 1206 | hCoV-19/Japan/PG-3344/2020\|EPI_ISL_691459\|2020-04 | .................... | .................... | ..................... | .................... | .................... | .................... |
| V | 1207 | hCoV-19/Japan/PG-3345/2020\|EPI_ISL_691460\|2020-04 | .................... | .................... | ..................... | .................... | .................... | .................... |
| V | 1208 | hCoV-19/Japan/PG-3346/2020\|EPI_ISL_691461\|2020-04 | .................... | .................... | ..................... | .................... | .................... | .................... |
| V | 1209 | hCoV-19/Japan/PG-3347/2020\|EPI_ISL_691462\|2020-04 | .................... | .................... | ..................... | .................... | .................... | .................... |
| V | 1210 | hCoV-19/Japan/PG-3348/2020\|EPI_ISL_691463\|2020-04 | .................... | .................... | ..................... | .................... | .................... | .................... |
| V | 1211 | hCoV-19/Japan/PG-3349/2020\|EPI_ISL_691464\|2020-04 | .................... | .................... | ..................... | .................... | .................... | .................... |
| V | 1212 | hCoV-19/Japan/PG-3350/2020\|EPI_ISL_691465\|2020-04 | .................... | .................... | ..................... | .................... | .................... | .................... |
| V | 1213 | hCoV-19/Japan/PG-3351/2020\|EPI_ISL_691466\|2020-04 | .................... | .................... | ..................... | .................... | .................... | .................... |
| V | 1214 | hCoV-19/Japan/PG-3352/2020\|EPI_ISL_691467\|2020-04 | .................... | .................... | ..................... | .................... | .................... | .................... |
| V | 1215 | hCoV-19/Japan/PG-3353/2020\|EPI_ISL_691468\|2020-04 | .................... | .................... | ..................... | .................... | .................... | .................... |
| V | 1216 | hCoV-19/Japan/PG-3354/2020\|EPI_ISL_691469\|2020-04 | .................... | .................... | ..................... | .................... | .................... | .................... |
| V | 1217 | hCoV-19/Japan/PG-3355/2020\|EPI_ISL_691470\|2020-04 | .................... | .................... | ..................... | .................... | .................... | .................... |
| V | 1218 | hCoV-19/Japan/PG-3356/2020\|EPI_ISL_691471\|2020-04 | .................... | .................... | ..................... | .................... | .................... | .................... |
| V | 1219 | hCoV-19/Japan/PG-3357/2020\|EPI_ISL_691472\|2020-04 | .................... | .................... | ..................... | .................... | .................... | .................... |
| V | 1220 | hCoV-19/Japan/PG-3358/2020\|EPI_ISL_691473\|2020-04 | .................... | .................... | ..................... | .................... | .................... | .................... |
| V | 1221 | hCoV-19/Japan/PG-3359/2020\|EPI_ISL_691474\|2020-04 | .................... | .................... | ..................... | .................... | .................... | .................... |
| V | 1222 | hCoV-19/Japan/PG-3360/2020\|EPI_ISL_691475\|2020-04 | .................... | .................... | ..................... | .................... | .................... | .................... |
| V | 1223 | hCoV-19/Japan/PG-3361/2020\|EPI_ISL_691476\|2020-04 | .................... | .................... | ..................... | .................... | .................... | .................... |
| V | 1224 | hCoV-19/Japan/PG-3362/2020\|EPI_ISL_691477\|2020-04 | .................... | .................... | ..................... | .................... | .................... | .................... |
| V | 1225 | hCoV-19/Japan/PG-3363/2020\|EPI_ISL_691478\|2020-04 | .................... | .................... | ..................... | .................... | .................... | .................... |
| V | 1226 | hCoV-19/Japan/PG-3364/2020\|EPI_ISL_691479\|2020-04 | .................... | .................... | ..................... | .................... | .................... | .................... |
| V | 1227 | hCoV-19/Japan/PG-3365/2020\|EPI_ISL_691480\|2020-04 | .................... | .................... | ..................... | .................... | .................... | .................... |
| V | 1228 | hCoV-19/Japan/PG-3366/2020\|EPI_ISL_691481\|2020-04 | .................... | .................... | ..................... | .................... | .................... | .................... |
| V | 1229 | hCoV-19/Japan/PG-3367/2020\|EPI_ISL_691482\|2020-04 | .................... | .................... | ..................... | .................... | .................... | .................... |
| V | 1230 | hCoV-19/Japan/PG-3368/2020\|EPI_ISL_691483\|2020-04 | .................... | .................... | ..................... | .................... | .................... | .................... |
| V | 1231 | hCoV-19/Japan/PG-4477/2020\|EPI_ISL_692700\|2020-07 | .................... | .................... | ..................... | .................... | .................... | .................... |
| V | 1232 | hCoV-19/Japan/PG-4478/2020\|EPI_ISL_692701\|2020-07 | .................... | .................... | ..................... | .................... | .................... | .................... |
| V | 1233 | hCoV-19/Japan/PG-4479/2020\|EPI_ISL_692702\|2020-07 | .................... | .................... | ..................... | .................... | .................... | .................... |
| V | 1234 | hCoV-19/Japan/PG-4480/2020\|EPI_ISL_692703\|2020-07 | .................... | .................... | ..................... | .................... | .................... | .................... |
| V | 1235 | hCoV-19/Japan/PG-4481/2020\|EPI_ISL_692704\|2020-07 | .................... | .................... | ..................... | .................... | .................... | .................... |
| V | 1236 | hCoV-19/Japan/PG-4482/2020\|EPI_ISL_692705\|2020-07 | .................... | .................... | ..................... | .................... | .................... | .................... |
| V | 1237 | hCoV-19/Japan/PG-4483/2020\|EPI_ISL_692706\|2020-07 | .................... | .................... | ..................... | .................... | .................... | .................... |
| V | 1238 | hCoV-19/Japan/PG-4484/2020\|EPI_ISL_692707\|2020-07 | .................... | .................... | ..................... | .................... | .................... | .................... |
| V | 1239 | hCoV-19/Japan/PG-4485/2020\|EPI_ISL_692708\|2020-07 | .................... | .................... | ..................... | .................... | .................... | .................... |
| V | 1240 | hCoV-19/Japan/PG-4486/2020\|EPI_ISL_692709\|2020-07 | .................... | .................... | ..................... | .................... | .................... | .................... |
| V | 1241 | hCoV-19/Japan/PG-4487/2020\|EPI_ISL_692710\|2020-07 | .................... | .................... | ..................... | .................... | .................... | .................... |
| V | 1242 | hCoV-19/Japan/PG-4488/2020\|EPI_ISL_692711\|2020-07 | .................... | .................... | ..................... | .................... | .................... | .................... |
| V | 1243 | hCoV-19/Japan/PG-4489/2020\|EPI_ISL_692712\|2020-07 | .................... | .................... | ..................... | .................... | .................... | .................... |
| V | 1244 | hCoV-19/Japan/PG-4490/2020\|EPI_ISL_692713\|2020-07 | .................... | .................... | ..................... | .................... | .................... | .................... |
| V | 1245 | hCoV-19/Japan/PG-4491/2020\|EPI_ISL_692714\|2020-07 | .................... | .................... | ..................... | .................... | .................... | .................... |
| V | 1246 | hCoV-19/Japan/PG-4492/2020\|EPI_ISL_692715\|2020-07 | .................... | .................... | ..................... | .................... | .................... | .................... |
| V | 1247 | hCoV-19/Japan/PG-4493/2020\|EPI_ISL_692716\|2020-07 | .................... | .................... | ..................... | .................... | .................... | .................... |

| | | | | | | | |
|---|---|---|---|---|---|---|---|
| V | 1248 hCoV-19/Japan/PG-4494/2020\|EPI_ISL_692717\|2020-07 | ···················· | ···················· | ····················· | ···················· | ···················· | ···················· |
| V | 1250 hCoV-19/Japan/PG-4496/2020\|EPI_ISL_692719\|2020-07 | ···················· | ···················· | ····················· | ···················· | ···················· | ···················· |
| V | 1251 hCoV-19/Japan/PG-4497/2020\|EPI_ISL_692720\|2020-07 | ···················· | ···················· | ····················· | ···················· | ···················· | ···················· |
| V | 1252 hCoV-19/Japan/PG-4498/2020\|EPI_ISL_692721\|2020-07 | ···················· | ···················· | ····················· | ···················· | ···················· | ···················· |
| V | 1253 hCoV-19/Japan/PG-4499/2020\|EPI_ISL_692722\|2020-07 | ···················· | ···················· | ····················· | ···················· | ···················· | ···················· |
| V | 1254 hCoV-19/Japan/PG-4500/2020\|EPI_ISL_692723\|2020-07 | ···················· | ···················· | ····················· | ···················· | ···················· | ···················· |
| V | 1255 hCoV-19/Japan/PG-4501/2020\|EPI_ISL_692724\|2020-07 | ···················· | ···················· | ····················· | ···················· | ···················· | ···················· |
| V | 1256 hCoV-19/Japan/PG-4502/2020\|EPI_ISL_692725\|2020-07 | ···················· | ···················· | ····················· | ···················· | ···················· | ···················· |
| V | 1257 hCoV-19/Japan/PG-4503/2020\|EPI_ISL_692726\|2020-07 | ···················· | ···················· | ····················· | ···················· | ···················· | ···················· |
| V | 1258 hCoV-19/Japan/PG-4504/2020\|EPI_ISL_692727\|2020-07 | ···················· | ···················· | ····················· | ···················· | ···················· | ···················· |
| V | 1259 hCoV-19/Japan/PG-4505/2020\|EPI_ISL_692728\|2020-07 | ···················· | ···················· | ····················· | ···················· | ···················· | ···················· |
| V | 1260 hCoV-19/Japan/PG-4506/2020\|EPI_ISL_692729\|2020-07 | ···················· | ···················· | ····················· | ···················· | ···················· | ···················· |
| V | 1261 hCoV-19/France/ARA-67612/2020\|EPI_ISL_692764\|2020- | ···················· | ···················· | ····················· | ···················· | ···················· | ···················· |
| V | 1262 hCoV-19/France/ARA-67686/2020\|EPI_ISL_692765\|2020- | ···················· | ···················· | ····················· | ···················· | ···················· | ···················· |
| V | 1263 hCoV-19/France/ARA-6777/2020\|EPI_ISL_692767\|2020-0 | ···················· | ···················· | ····················· | ···················· | ···················· | ···················· |
| V | 1264 hCoV-19/USA/MA-MASPHL-01089/2020\|EPI_ISL_692770\|20 | ···················· | ···················· | ····················· | ···················· | ···················· | ···················· |
| V | 1265 hCoV-19/USA/MA-MASPHL-01088/2020\|EPI_ISL_692771\|20 | ···················· | ···················· | ····················· | ···················· | ···················· | ···················· |
| V | 1266 hCoV-19/USA/MA-MASPHL-01086/2020\|EPI_ISL_692772\|20 | ···················· | ···················· | ····················· | ···················· | ···················· | ···················· |
| V | 1267 hCoV-19/USA/MA-MASPHL-01085/2020\|EPI_ISL_692773\|20 | ···················· | ···················· | ····················· | ···················· | ···················· | ···················· |
| V | 1268 hCoV-19/USA/MA-MASPHL-01084/2020\|EPI_ISL_692774\|20 | ···················· | ···················· | ····················· | ···················· | ···················· | ···················· |
| V | 1269 hCoV-19/USA/MA-MASPHL-01061/2020\|EPI_ISL_692776\|20 | ···················· | ···················· | ····················· | ···················· | ···················· | ···················· |
| V | 1270 hCoV-19/USA/MA-MASPHL-01035/2020\|EPI_ISL_692778\|20 | ···················· | ···················· | ····················· | ···················· | ···················· | ···················· |
| V | 1271 hCoV-19/USA/MA-MASPHL-01032/2020\|EPI_ISL_692780\|20 | ···················· | ···················· | ····················· | ···················· | ···················· | ···················· |
| V | 1272 hCoV-19/USA/MA-MASPHL-01029/2020\|EPI_ISL_692782\|20 | ···················· | ···················· | ····················· | ···················· | ···················· | ···················· |
| V | 1273 hCoV-19/USA/MA-MASPHL-01024/2020\|EPI_ISL_692785\|20 | ···················· | ·A·················· | ····················· | ···················· | ···················· | ···················· |
| V | 1274 hCoV-19/USA/MA-MASPHL-01021/2020\|EPI_ISL_692787\|20 | ···················· | ···················· | ····················· | ···················· | ···················· | ···················· |
| V | 1275 hCoV-19/USA/MA-MASPHL-01020/2020\|EPI_ISL_692788\|20 | ···················· | ···················· | ····················· | ···················· | ···················· | ···················· |
| V | 1276 hCoV-19/USA/MA-MASPHL-01019/2020\|EPI_ISL_692789\|20 | ···················· | ···················· | ····················· | ···················· | ···················· | ···················· |
| V | 1277 hCoV-19/USA/MA-MASPHL-01016/2020\|EPI_ISL_692792\|20 | ···················· | ···················· | ····················· | ···················· | ···················· | ···················· |
| V | 1278 hCoV-19/USA/MA-MASPHL-01014/2020\|EPI_ISL_692794\|20 | ···················· | ···················· | ····················· | ···················· | ···················· | ···················· |
| V | 1279 hCoV-19/USA/MA-MASPHL-01013/2020\|EPI_ISL_692795\|20 | ···················· | ·A·················· | ····················· | ···················· | ···················· | ···················· |
| V | 1280 hCoV-19/USA/MA-MASPHL-01010/2020\|EPI_ISL_692796\|20 | ···················· | ···················· | ····················· | ···················· | ···················· | ···················· |
| V | 1281 hCoV-19/Japan/PG-0629/2020\|EPI_ISL_684239\|2020-04 | ···················· | ···················· | ····················· | ···················· | ···················· | ···················· |
| V | 1282 hCoV-19/Japan/PG-0630/2020\|EPI_ISL_684240\|2020-04 | ···················· | ···················· | ····················· | ···················· | ···················· | ···················· |
| V | 1283 hCoV-19/Japan/PG-0631/2020\|EPI_ISL_684241\|2020-04 | ···················· | ···················· | ····················· | ···················· | ···················· | ···················· |
| V | 1284 hCoV-19/Japan/PG-0632/2020\|EPI_ISL_684242\|2020-04 | ···················· | ···················· | ····················· | ···················· | ···················· | ···················· |
| V | 1285 hCoV-19/Japan/PG-0633/2020\|EPI_ISL_684243\|2020-04 | ···················· | ···················· | ····················· | ···················· | ···················· | ···················· |
| V | 1286 hCoV-19/Japan/PG-0634/2020\|EPI_ISL_684244\|2020-04 | ···················· | ···················· | ····················· | ···················· | ···················· | ···················· |
| V | 1287 hCoV-19/Japan/PG-0635/2020\|EPI_ISL_684245\|2020-04 | ···················· | ···················· | ····················· | ···················· | ···················· | ···················· |
| V | 1288 hCoV-19/Japan/PG-0636/2020\|EPI_ISL_684246\|2020-04 | ···················· | ···················· | ····················· | ···················· | ···················· | ···················· |
| V | 1289 hCoV-19/Japan/PG-0637/2020\|EPI_ISL_684247\|2020-04 | ···················· | ···················· | ····················· | ···················· | ···················· | ···················· |
| V | 1290 hCoV-19/Japan/PG-0638/2020\|EPI_ISL_684248\|2020-04 | ···················· | ···················· | ····················· | ···················· | ···················· | ···················· |
| V | 1291 hCoV-19/Japan/PG-0639/2020\|EPI_ISL_684249\|2020-04 | ···················· | ···················· | ····················· | ···················· | ···················· | ···················· |
| V | 1292 hCoV-19/Japan/PG-0640/2020\|EPI_ISL_684250\|2020-04 | ···················· | ···················· | ····················· | ···················· | ···················· | ···················· |
| V | 1293 hCoV-19/Japan/PG-0641/2020\|EPI_ISL_684251\|2020-04 | ···················· | ···················· | ····················· | ···················· | ···················· | ···················· |
| V | 1294 hCoV-19/Japan/PG-0642/2020\|EPI_ISL_684252\|2020-04 | ···················· | ···················· | ····················· | ···················· | ···················· | ···················· |
| V | 1295 hCoV-19/Japan/PG-0643/2020\|EPI_ISL_684253\|2020-04 | ···················· | ···················· | ····················· | ···················· | ···················· | ···················· |
| V | 1296 hCoV-19/Japan/PG-0644/2020\|EPI_ISL_684254\|2020-04 | ···················· | ···················· | ····················· | ···················· | ···················· | ···················· |
| V | 1297 hCoV-19/Japan/PG-0645/2020\|EPI_ISL_684255\|2020-04 | ···················· | ···················· | ····················· | ···················· | ···················· | ···················· |
| V | 1298 hCoV-19/Japan/PG-0646/2020\|EPI_ISL_684256\|2020-04 | ···················· | ···················· | ····················· | ···················· | ···················· | ···················· |
| V | 1299 hCoV-19/Japan/PG-0647/2020\|EPI_ISL_684257\|2020-04 | ···················· | ···················· | ····················· | ···················· | ···················· | ···················· |
| V | 1300 hCoV-19/Japan/PG-0648/2020\|EPI_ISL_684258\|2020-04 | ···················· | ···················· | ····················· | ···················· | ···················· | ···················· |
| V | 1301 hCoV-19/Japan/PG-0649/2020\|EPI_ISL_684259\|2020-04 | ···················· | ···················· | ····················· | ···················· | ···················· | ···················· |
| V | 1302 hCoV-19/Japan/PG-0650/2020\|EPI_ISL_684260\|2020-04 | ···················· | ···················· | ····················· | ···················· | ···················· | ···················· |
| V | 1303 hCoV-19/Japan/PG-0651/2020\|EPI_ISL_684261\|2020-04 | ···················· | ···················· | ····················· | ···················· | ···················· | ···················· |
| V | 1304 hCoV-19/Japan/PG-0652/2020\|EPI_ISL_684262\|2020-04 | ···················· | ···················· | ····················· | ···················· | ···················· | ···················· |
| V | 1305 hCoV-19/Japan/PG-0653/2020\|EPI_ISL_684263\|2020-04 | ···················· | ···················· | ····················· | ···················· | ···················· | ···················· |
| V | 1306 hCoV-19/Japan/PG-0654/2020\|EPI_ISL_684264\|2020-04 | ···················· | ···················· | ····················· | ···················· | ···················· | ···················· |
| V | 1307 hCoV-19/Japan/PG-0655/2020\|EPI_ISL_684265\|2020-04 | ···················· | ···················· | ····················· | ···················· | ···················· | ···················· |
| V | 1308 hCoV-19/Japan/PG-0656/2020\|EPI_ISL_684266\|2020-04 | ···················· | ···················· | ····················· | ···················· | ···················· | ···················· |
| V | 1309 hCoV-19/Japan/PG-0657/2020\|EPI_ISL_684267\|2020-04 | ···················· | ···················· | ····················· | ···················· | ···················· | ···················· |
| V | 1310 hCoV-19/Japan/PG-0658/2020\|EPI_ISL_684268\|2020-04 | ···················· | ···················· | ····················· | ···················· | ···················· | ···················· |
| V | 1311 hCoV-19/Japan/PG-0659/2020\|EPI_ISL_684269\|2020-04 | ···················· | ···················· | ····················· | ···················· | ···················· | ···················· |
| V | 1312 hCoV-19/Japan/PG-0660/2020\|EPI_ISL_684270\|2020-04 | ···················· | ···················· | ····················· | ···················· | ···················· | ···················· |
| V | 1313 hCoV-19/Japan/PG-0661/2020\|EPI_ISL_684271\|2020-04 | ···················· | ···················· | ····················· | ···················· | ···················· | ···················· |
| V | 1314 hCoV-19/Japan/PG-0662/2020\|EPI_ISL_684272\|2020-04 | ···················· | ···················· | ····················· | ···················· | ···················· | ···················· |
| V | 1315 hCoV-19/Japan/PG-0663/2020\|EPI_ISL_684273\|2020-04 | ···················· | ···················· | ····················· | ···················· | ···················· | ···················· |
| V | 1316 hCoV-19/Japan/PG-0664/2020\|EPI_ISL_684274\|2020-04 | ···················· | ···················· | ····················· | ···················· | ···················· | ···················· |
| V | 1317 hCoV-19/Japan/PG-0665/2020\|EPI_ISL_684275\|2020-04 | ···················· | ···················· | ····················· | ···················· | ···················· | ···················· |
| V | 1318 hCoV-19/Japan/PG-0666/2020\|EPI_ISL_684276\|2020-04 | ···················· | ···················· | ····················· | ···················· | ···················· | ···················· |
| V | 1319 hCoV-19/Japan/PG-0667/2020\|EPI_ISL_684277\|2020-04 | ···················· | ···················· | ····················· | ···················· | ···················· | ···················· |
| V | 1320 hCoV-19/Japan/PG-0668/2020\|EPI_ISL_684278\|2020-04 | ···················· | ···················· | ····················· | ···················· | ···················· | ···················· |
| V | 1321 hCoV-19/Japan/PG-1894/2020\|EPI_ISL_685500\|2020-04 | ···················· | ···················· | ····················· | ···················· | ···················· | ···················· |
| V | 1322 hCoV-19/Japan/PG-1897/2020\|EPI_ISL_685503\|2020-04 | ···················· | ···················· | ····················· | ···················· | ···················· | ···················· |
| V | 1323 hCoV-19/Japan/PG-1898/2020\|EPI_ISL_685504\|2020-04 | ···················· | ···················· | ····················· | ···················· | ···················· | ···················· |
| V | 1324 hCoV-19/Japan/PG-1899/2020\|EPI_ISL_685505\|2020-05 | ···················· | ···················· | ····················· | ···················· | ···················· | ···················· |
| V | 1325 hCoV-19/Japan/PG-1900/2020\|EPI_ISL_685506\|2020-04 | ···················· | ···················· | ····················· | ···················· | ···················· | ···················· |
| V | 1326 hCoV-19/Japan/PG-1901/2020\|EPI_ISL_685507\|2020-04 | ···················· | ···················· | ····················· | ···················· | ···················· | ···················· |
| V | 1327 hCoV-19/Japan/PG-1902/2020\|EPI_ISL_685508\|2020-04 | ···················· | ···················· | ····················· | ···················· | ···················· | ···················· |
| V | 1328 hCoV-19/Japan/PG-1903/2020\|EPI_ISL_685509\|2020-04 | ···················· | ···················· | ····················· | ···················· | ···················· | ···················· |

| | | | | | | | |
|---|---|---|---|---|---|---|---|
| V | 1329 hCoV-19/Japan/PG-1904/2020\|EPI_ISL_685510\|2020-04 | .................... | .................... | .................... | .................... | .................... | .................... |
| V | 1330 hCoV-19/Japan/PG-1964/2020\|EPI_ISL_685570\|2020-04 | .................... | .................... | .................... | .................... | .................... | .................... |
| V | 1331 hCoV-19/Japan/PG-1965/2020\|EPI_ISL_685571\|2020-04 | .................... | .................... | .................... | .................... | .................... | .................... |
| V | 1332 hCoV-19/Japan/PG-1966/2020\|EPI_ISL_685572\|2020-04 | .................... | .................... | .................... | .................... | .................... | .................... |
| V | 1333 hCoV-19/Japan/PG-1967/2020\|EPI_ISL_685573\|2020-04 | .................... | .................... | .................... | .................... | .................... | .................... |
| V | 1334 hCoV-19/Japan/PG-1968/2020\|EPI_ISL_685574\|2020-05 | .................... | .................... | .................... | .................... | .................... | .................... |
| V | 1335 hCoV-19/Japan/PG-1969/2020\|EPI_ISL_685575\|2020-05 | .................... | .................... | .................... | .................... | .................... | .................... |
| V | 1336 hCoV-19/Japan/PG-1970/2020\|EPI_ISL_685576\|2020-05 | .................... | .................... | .................... | .................... | .................... | .................... |
| V | 1337 hCoV-19/Japan/PG-1971/2020\|EPI_ISL_685577\|2020-05 | .................... | .................... | .................... | .................... | .................... | .................... |
| V | 1338 hCoV-19/Japan/PG-1972/2020\|EPI_ISL_685578\|2020-05 | .................... | .................... | .................... | .................... | .................... | .................... |
| V | 1339 hCoV-19/Japan/PG-1973/2020\|EPI_ISL_685579\|2020-05 | .................... | .................... | .................... | .................... | .................... | .................... |
| V | 1340 hCoV-19/Japan/PG-1974/2020\|EPI_ISL_685580\|2020-05 | .................... | .................... | .................... | .................... | .................... | .................... |
| V | 1341 hCoV-19/Japan/PG-1975/2020\|EPI_ISL_685581\|2020-05 | .................... | .................... | .................... | .................... | .................... | .................... |
| V | 1342 hCoV-19/Japan/PG-1976/2020\|EPI_ISL_685582\|2020-05 | .................... | .................... | .................... | .................... | .................... | .................... |
| V | 1343 hCoV-19/Japan/PG-1977/2020\|EPI_ISL_685583\|2020-05 | .................... | .................... | .................... | .................... | .................... | .................... |
| V | 1345 hCoV-19/Japan/PG-1980/2020\|EPI_ISL_685586\|2020-03 | .................... | .................... | .................... | .................... | .................... | .................... |
| V | 1346 hCoV-19/Japan/PG-1981/2020\|EPI_ISL_685587\|2020-04 | .................... | .................... | .................... | .................... | .................... | .................... |
| V | 1347 hCoV-19/Japan/PG-1982/2020\|EPI_ISL_685588\|2020-04 | .................... | .................... | .................... | .................... | .................... | .................... |
| V | 1348 hCoV-19/Japan/PG-1983/2020\|EPI_ISL_685589\|2020-04 | .................... | .................... | .................... | .................... | .................... | .................... |
| V | 1349 hCoV-19/Japan/PG-1984/2020\|EPI_ISL_685590\|2020-04 | .................... | .................... | .................... | .................... | .................... | .................... |
| V | 1350 hCoV-19/Japan/PG-1985/2020\|EPI_ISL_685591\|2020-04 | .................... | .................... | .................... | .................... | .................... | .................... |
| V | 1351 hCoV-19/Japan/PG-1986/2020\|EPI_ISL_685592\|2020-04 | .................... | .................... | .................... | .................... | .................... | .................... |
| V | 1352 hCoV-19/Japan/PG-1987/2020\|EPI_ISL_685593\|2020-04 | .................... | .................... | .................... | .................... | .................... | .................... |
| V | 1353 hCoV-19/Japan/PG-1989/2020\|EPI_ISL_685594\|2020-04 | .................... | .................... | .................... | .................... | .................... | .................... |
| V | 1354 hCoV-19/Japan/PG-1990/2020\|EPI_ISL_685595\|2020-04 | .................... | .................... | .................... | .................... | .................... | .................... |
| V | 1355 hCoV-19/Japan/PG-1991/2020\|EPI_ISL_685596\|2020-04 | .................... | .................... | .................... | .................... | .................... | .................... |
| V | 1356 hCoV-19/Japan/PG-1992/2020\|EPI_ISL_685597\|2020-04 | .................... | .................... | .................... | .................... | .................... | .................... |
| V | 1357 hCoV-19/Japan/PG-1993/2020\|EPI_ISL_685598\|2020-04 | .................... | .................... | .................... | .................... | .................... | .................... |
| V | 1358 hCoV-19/Japan/PG-1994/2020\|EPI_ISL_685599\|2020-04 | .................... | .................... | .................... | .................... | .................... | .................... |
| V | 1359 hCoV-19/Japan/PG-7698/2020\|EPI_ISL_686800\|2020-08 | .................... | .................... | .................... | .................... | .................... | .................... |
| V | 1360 hCoV-19/Japan/PG-7699/2020\|EPI_ISL_686801\|2020-08 | .................... | .................... | .................... | .................... | .................... | .................... |
| V | 1361 hCoV-19/Japan/PG-7700/2020\|EPI_ISL_686802\|2020-08 | .................... | .................... | .................... | .................... | .................... | .................... |
| V | 1362 hCoV-19/Japan/PG-7701/2020\|EPI_ISL_686803\|2020-08 | .................... | .................... | .................... | .................... | .................... | .................... |
| V | 1363 hCoV-19/Japan/PG-7702/2020\|EPI_ISL_686804\|2020-08 | .................... | .................... | .................... | .................... | .................... | .................... |
| V | 1364 hCoV-19/Japan/PG-7703/2020\|EPI_ISL_686805\|2020-08 | .................... | .................... | .................... | .................... | .................... | .................... |
| V | 1365 hCoV-19/Japan/PG-7704/2020\|EPI_ISL_686806\|2020-08 | .................... | .................... | .................... | .................... | .................... | .................... |
| V | 1366 hCoV-19/Japan/PG-7705/2020\|EPI_ISL_686807\|2020-08 | .................... | .................... | .................... | .................... | .................... | .................... |
| V | 1367 hCoV-19/Japan/PG-7706/2020\|EPI_ISL_686808\|2020-08 | .................... | .................... | .................... | .................... | .................... | .................... |
| V | 1368 hCoV-19/Japan/PG-7707/2020\|EPI_ISL_686809\|2020-08 | .................... | .................... | .................... | .................... | .................... | .................... |
| V | 1369 hCoV-19/Japan/PG-7708/2020\|EPI_ISL_686810\|2020-08 | .................... | .................... | .................... | .................... | .................... | .................... |
| V | 1370 hCoV-19/Japan/PG-7709/2020\|EPI_ISL_686811\|2020-08 | .................... | .................... | .................... | .................... | .................... | .................... |
| V | 1371 hCoV-19/Japan/PG-7710/2020\|EPI_ISL_686812\|2020-08 | .................... | .................... | .................... | .................... | .................... | .................... |
| V | 1372 hCoV-19/Japan/PG-7711/2020\|EPI_ISL_686813\|2020-08 | .................... | .................... | .................... | .................... | .................... | .................... |
| V | 1373 hCoV-19/Japan/PG-7712/2020\|EPI_ISL_686814\|2020-08 | .................... | .................... | .................... | .................... | .................... | .................... |
| V | 1374 hCoV-19/Japan/PG-7713/2020\|EPI_ISL_686815\|2020-08 | .................... | .................... | .................... | .................... | .................... | .................... |
| V | 1375 hCoV-19/Japan/PG-7714/2020\|EPI_ISL_686816\|2020-08 | .................... | .................... | .................... | .................... | .................... | .................... |
| V | 1376 hCoV-19/Japan/PG-7715/2020\|EPI_ISL_686817\|2020-08 | .................... | .................... | .................... | .................... | .................... | .................... |
| V | 1377 hCoV-19/Japan/PG-7716/2020\|EPI_ISL_686818\|2020-08 | .................... | .................... | .................... | .................... | .................... | .................... |
| V | 1378 hCoV-19/Japan/PG-7717/2020\|EPI_ISL_686819\|2020-08 | .................... | .................... | .................... | .................... | .................... | .................... |
| V | 1379 hCoV-19/Japan/PG-7718/2020\|EPI_ISL_686820\|2020-08 | ........T........... | .................... | .................... | .................... | .................... | .................... |
| V | 1380 hCoV-19/Japan/PG-7719/2020\|EPI_ISL_686821\|2020-08 | .................... | .................... | .................... | .................... | .................... | .................... |
| V | 1381 hCoV-19/Japan/PG-7720/2020\|EPI_ISL_686822\|2020-08 | .................... | .................... | .................... | .................... | .................... | .................... |
| V | 1382 hCoV-19/Japan/PG-7721/2020\|EPI_ISL_686823\|2020-08 | .................... | .................... | .................... | .................... | .................... | .................... |
| V | 1383 hCoV-19/Japan/PG-7722/2020\|EPI_ISL_686824\|2020-08 | .................... | .................... | .................... | .................... | .................... | .................... |
| V | 1384 hCoV-19/Japan/PG-7723/2020\|EPI_ISL_686825\|2020-08 | .................... | .................... | .................... | .................... | .................... | .................... |
| V | 1385 hCoV-19/Japan/PG-7724/2020\|EPI_ISL_686826\|2020-08 | .................... | .................... | .................... | .................... | .................... | .................... |
| V | 1386 hCoV-19/Japan/PG-7725/2020\|EPI_ISL_686827\|2020-08 | .................... | .................... | .................... | .................... | .................... | .................... |
| V | 1387 hCoV-19/Japan/PG-7726/2020\|EPI_ISL_686828\|2020-08 | .................... | .................... | .................... | .................... | .................... | .................... |
| V | 1388 hCoV-19/Japan/PG-7727/2020\|EPI_ISL_686829\|2020-08 | .................... | .................... | .................... | .................... | .................... | .................... |
| V | 1389 hCoV-19/Japan/PG-7728/2020\|EPI_ISL_686830\|2020-08 | .................... | .................... | .................... | .................... | .................... | .................... |
| V | 1390 hCoV-19/Japan/PG-7729/2020\|EPI_ISL_686831\|2020-09 | .................... | .................... | .................... | .................... | .................... | .................... |
| V | 1391 hCoV-19/Japan/PG-7730/2020\|EPI_ISL_686832\|2020-09 | .................... | .................... | .................... | .................... | .................... | .................... |
| V | 1392 hCoV-19/Japan/PG-7731/2020\|EPI_ISL_686833\|2020-09 | .................... | .................... | .................... | .................... | .................... | .................... |
| V | 1393 hCoV-19/Japan/PG-7732/2020\|EPI_ISL_686834\|2020-09 | .................... | .................... | .................... | .................... | .................... | .................... |
| V | 1394 hCoV-19/Japan/PG-7733/2020\|EPI_ISL_686835\|2020-09 | .................... | .................... | .................... | .................... | .................... | .................... |
| V | 1395 hCoV-19/Japan/PG-7734/2020\|EPI_ISL_686836\|2020-09 | .................... | .................... | .................... | .................... | .................... | .................... |
| V | 1396 hCoV-19/Japan/PG-7735/2020\|EPI_ISL_686837\|2020-09 | .................... | .................... | .................... | .................... | .................... | .................... |
| V | 1397 hCoV-19/Japan/PG-7736/2020\|EPI_ISL_686838\|2020-09 | .................... | .................... | .................... | .................... | .................... | .................... |
| V | 1398 hCoV-19/Japan/PG-7737/2020\|EPI_ISL_686839\|2020-09 | .................... | .................... | .................... | .................... | .................... | .................... |
| V | 1399 hCoV-19/Japan/PG-7739/2020\|EPI_ISL_686841\|2020-09 | .................... | .................... | .................... | .................... | .................... | .................... |
| V | 1400 hCoV-19/Japan/PG-5935/2020\|EPI_ISL_689050\|2020 | .................... | .................... | .................... | .................... | .................... | .................... |
| V | 1401 hCoV-19/Japan/PG-5936/2020\|EPI_ISL_689051\|2020 | .................... | .................... | .................... | .................... | .................... | .................... |
| V | 1402 hCoV-19/Japan/PG-5937/2020\|EPI_ISL_689052\|2020 | .................... | .................... | .................... | .................... | .................... | .................... |
| V | 1403 hCoV-19/Japan/PG-5938/2020\|EPI_ISL_689053\|2020 | .................... | .................... | .................... | .................... | .................... | .................... |
| V | 1404 hCoV-19/Japan/PG-5939/2020\|EPI_ISL_689054\|2020 | .................... | .................... | .................... | .................... | .................... | .................... |
| V | 1405 hCoV-19/Japan/PG-5940/2020\|EPI_ISL_689055\|2020 | .................... | .................... | .................... | .................... | .................... | .................... |
| V | 1406 hCoV-19/Japan/PG-5941/2020\|EPI_ISL_689056\|2020 | .................... | .................... | .................... | .................... | .................... | .................... |
| V | 1407 hCoV-19/Japan/PG-5942/2020\|EPI_ISL_689057\|2020 | .................... | .................... | .................... | .................... | .................... | .................... |
| V | 1408 hCoV-19/Japan/PG-5943/2020\|EPI_ISL_689058\|2020 | .................... | .................... | .................... | .................... | .................... | .................... |
| V | 1409 hCoV-19/Japan/PG-5944/2020\|EPI_ISL_689059\|2020 | .................... | .................... | .................... | .................... | .................... | .................... |

| | | | | | | | | |
|---|---|---|---|---|---|---|---|---|
| V | 1410 | hCoV-19/Japan/PG-5945/2020\|EPI_ISL_689060\|2020 | .................... | .................... | ..................... | .................... | .................... | .................... |
| V | 1411 | hCoV-19/Japan/PG-5946/2020\|EPI_ISL_689061\|2020 | .................... | .................... | ..................... | .................... | .................... | .................... |
| V | 1412 | hCoV-19/Japan/PG-5947/2020\|EPI_ISL_689062\|2020 | .................... | .................... | ..................... | .................... | .................... | .................... |
| V | 1413 | hCoV-19/Japan/PG-5948/2020\|EPI_ISL_689063\|2020 | .................... | .................... | ..................... | .................... | .................... | .................... |
| V | 1414 | hCoV-19/Japan/PG-5949/2020\|EPI_ISL_689064\|2020 | .................... | .................... | ..................... | .................... | .................... | .................... |
| V | 1415 | hCoV-19/Japan/PG-5950/2020\|EPI_ISL_689065\|2020 | .................... | .................... | ..................... | .................... | .................... | .................... |
| V | 1416 | hCoV-19/Japan/PG-5951/2020\|EPI_ISL_689066\|2020 | .................... | .................... | ..................... | .................... | .................... | .................... |
| V | 1417 | hCoV-19/Japan/PG-5952/2020\|EPI_ISL_689067\|2020 | .................... | .................... | ..................... | .................... | .................... | .................... |
| V | 1418 | hCoV-19/Japan/PG-5953/2020\|EPI_ISL_689068\|2020 | .................... | .................... | ..................... | .................... | .................... | .................... |
| V | 1419 | hCoV-19/Japan/PG-5954/2020\|EPI_ISL_689069\|2020 | .................... | .................... | ..................... | .................... | .................... | .................... |
| V | 1420 | hCoV-19/Japan/PG-5955/2020\|EPI_ISL_689070\|2020 | .................... | .................... | ..................... | .................... | .................... | .................... |
| V | 1421 | hCoV-19/Japan/PG-5956/2020\|EPI_ISL_689071\|2020 | .................... | .................... | ..................... | .................... | .................... | .................... |
| V | 1422 | hCoV-19/Japan/PG-5957/2020\|EPI_ISL_689072\|2020 | .................... | .................... | ..................... | .................... | .................... | .................... |
| V | 1423 | hCoV-19/Japan/PG-5958/2020\|EPI_ISL_689073\|2020 | .................... | .................... | ..................... | .................... | .................... | .................... |
| V | 1424 | hCoV-19/Japan/PG-5959/2020\|EPI_ISL_689074\|2020 | .................... | .................... | ..................... | .................... | .................... | .................... |
| V | 1425 | hCoV-19/Japan/PG-5960/2020\|EPI_ISL_689075\|2020 | .................... | .................... | ..................... | .................... | .................... | .................... |
| V | 1426 | hCoV-19/Japan/PG-5961/2020\|EPI_ISL_689076\|2020 | .................... | .................... | ..................... | .................... | .................... | .................... |
| V | 1427 | hCoV-19/Japan/PG-5962/2020\|EPI_ISL_689077\|2020-07 | .................... | .................... | ..................... | .................... | .................... | .................... |
| V | 1428 | hCoV-19/Japan/PG-5963/2020\|EPI_ISL_689078\|2020-07 | .................... | .................... | ..................... | .................... | .................... | .................... |
| V | 1429 | hCoV-19/Japan/PG-5964/2020\|EPI_ISL_689079\|2020-07 | .................... | .................... | ..................... | .................... | .................... | .................... |
| V | 1430 | hCoV-19/Japan/PG-5965/2020\|EPI_ISL_689080\|2020-07 | .................... | .................... | ..................... | .................... | .................... | .................... |
| V | 1431 | hCoV-19/Japan/PG-5966/2020\|EPI_ISL_689081\|2020-07 | .................... | .................... | ..................... | .................... | .................... | .................... |
| V | 1432 | hCoV-19/Japan/PG-5967/2020\|EPI_ISL_689082\|2020-07 | .................... | .................... | ..................... | .................... | .................... | .................... |
| V | 1433 | hCoV-19/Japan/PG-5968/2020\|EPI_ISL_689083\|2020-07 | .................... | .................... | ..................... | .................... | .................... | .................... |
| V | 1434 | hCoV-19/Japan/PG-5969/2020\|EPI_ISL_689084\|2020-07 | .................... | .................... | ..................... | .................... | .................... | .................... |
| V | 1435 | hCoV-19/Japan/PG-5970/2020\|EPI_ISL_689085\|2020-07 | .................... | .................... | ..................... | .................... | .................... | .................... |
| V | 1436 | hCoV-19/Japan/PG-5971/2020\|EPI_ISL_689086\|2020-07 | .................... | .................... | ..................... | .................... | .................... | .................... |
| V | 1437 | hCoV-19/Japan/PG-5972/2020\|EPI_ISL_689087\|2020-07 | .................... | .................... | ..................... | .................... | .................... | .................... |
| V | 1438 | hCoV-19/Japan/PG-5973/2020\|EPI_ISL_689088\|2020-07 | .................... | .................... | ..................... | .................... | .................... | .................... |
| V | 1439 | hCoV-19/Japan/PG-5974/2020\|EPI_ISL_689089\|2020-08 | .................... | .................... | ..................... | .................... | .................... | .................... |
| V | 1440 | hCoV-19/Japan/PG-5975/2020\|EPI_ISL_689090\|2020-08 | .................... | .................... | ..................... | .................... | .................... | .................... |
| V | 1441 | hCoV-19/Japan/PG-5976/2020\|EPI_ISL_689091\|2020-08 | .................... | .................... | ..................... | .................... | .................... | .................... |
| V | 1442 | hCoV-19/Japan/PG-5977/2020\|EPI_ISL_689092\|2020-08 | .................... | .................... | ..................... | .................... | .................... | .................... |
| V | 1443 | hCoV-19/Japan/PG-5978/2020\|EPI_ISL_689093\|2020-08 | .................... | .................... | ..................... | .................... | .................... | .................... |
| V | 1444 | hCoV-19/Japan/PG-5979/2020\|EPI_ISL_689094\|2020-08 | .................... | .................... | ..................... | .................... | .................... | .................... |
| V | 1445 | hCoV-19/Japan/PG-5980/2020\|EPI_ISL_689095\|2020-08 | .................... | .................... | ..................... | .................... | .................... | .................... |
| V | 1446 | hCoV-19/Japan/PG-5981/2020\|EPI_ISL_689096\|2020-08 | .................... | .................... | ..................... | .................... | .................... | .................... |
| V | 1447 | hCoV-19/Japan/PG-5982/2020\|EPI_ISL_689097\|2020-08 | .................... | .................... | ..................... | .................... | .................... | .................... |
| V | 1448 | hCoV-19/Japan/PG-5983/2020\|EPI_ISL_689098\|2020-08 | .................... | .................... | ..................... | .................... | .................... | .................... |
| V | 1449 | hCoV-19/Japan/PG-5986/2020\|EPI_ISL_689099\|2020-08 | .................... | .................... | ..................... | .................... | .................... | .................... |
| V | 1450 | hCoV-19/Japan/PG-4912/2020\|EPI_ISL_690032\|2020-07 | .................... | .................... | ..................... | .................... | .................... | .................... |
| V | 1451 | hCoV-19/Japan/PG-4913/2020\|EPI_ISL_690033\|2020-07 | .................... | .................... | ..................... | .................... | .................... | .................... |
| V | 1452 | hCoV-19/Japan/PG-4914/2020\|EPI_ISL_690034\|2020-07 | .................... | .................... | ..................... | .................... | .................... | .................... |
| V | 1453 | hCoV-19/Japan/PG-4915/2020\|EPI_ISL_690035\|2020-07 | .................... | .................... | ..................... | .................... | .................... | .................... |
| V | 1454 | hCoV-19/Japan/PG-4916/2020\|EPI_ISL_690036\|2020-07 | .................... | .................... | ..................... | .................... | .................... | .................... |
| V | 1455 | hCoV-19/Japan/PG-4917/2020\|EPI_ISL_690037\|2020-08 | .................... | .................... | ..................... | .................... | .................... | .................... |
| V | 1456 | hCoV-19/Japan/PG-4918/2020\|EPI_ISL_690038\|2020-08 | .................... | .................... | ..................... | .................... | .................... | .................... |
| V | 1458 | hCoV-19/Japan/PG-4920/2020\|EPI_ISL_690040\|2020-08 | .................... | .................... | ..................... | .................... | .................... | .................... |
| V | 1459 | hCoV-19/Japan/PG-4921/2020\|EPI_ISL_690041\|2020-08 | .................... | .................... | ..................... | .................... | .................... | .................... |
| V | 1460 | hCoV-19/Japan/PG-4922/2020\|EPI_ISL_690042\|2020-08 | .................... | .................... | ..................... | .................... | .................... | .................... |
| V | 1461 | hCoV-19/Japan/PG-4923/2020\|EPI_ISL_690043\|2020-08 | .................... | .................... | ..................... | .................... | .................... | .................... |
| V | 1462 | hCoV-19/Japan/PG-4924/2020\|EPI_ISL_690044\|2020-07 | .................... | .................... | ..................... | .................... | .................... | .................... |
| V | 1463 | hCoV-19/Japan/PG-4925/2020\|EPI_ISL_690045\|2020-07 | .................... | .................... | ..................... | .................... | .................... | .................... |
| V | 1464 | hCoV-19/Japan/PG-4926/2020\|EPI_ISL_690046\|2020-07 | .................... | .................... | ..................... | .................... | .................... | .................... |
| V | 1465 | hCoV-19/Japan/PG-4927/2020\|EPI_ISL_690047\|2020-07 | .................... | .................... | ..................... | .................... | .................... | .................... |
| V | 1466 | hCoV-19/Japan/PG-4928/2020\|EPI_ISL_690048\|2020-07 | .................... | .................... | ..................... | .................... | .................... | .................... |
| V | 1467 | hCoV-19/Japan/PG-4929/2020\|EPI_ISL_690049\|2020-07 | .................... | .................... | ..................... | .................... | .................... | .................... |
| V | 1468 | hCoV-19/Japan/PG-4930/2020\|EPI_ISL_690050\|2020-07 | .................... | .................... | ..................... | .................... | .................... | .................... |
| V | 1469 | hCoV-19/Japan/PG-4931/2020\|EPI_ISL_690051\|2020-07 | .................... | .................... | ..................... | .................... | .................... | .................... |
| V | 1470 | hCoV-19/Japan/PG-4932/2020\|EPI_ISL_690052\|2020-07 | .................... | .................... | ..................... | .................... | .................... | .................... |
| V | 1471 | hCoV-19/Japan/PG-4933/2020\|EPI_ISL_690053\|2020-07 | .................... | .................... | ..................... | .................... | .................... | .................... |
| V | 1472 | hCoV-19/Japan/PG-4934/2020\|EPI_ISL_690054\|2020-07 | .................... | .................... | ..................... | .................... | .................... | .................... |
| V | 1473 | hCoV-19/Japan/PG-4935/2020\|EPI_ISL_690055\|2020-08 | .................... | .................... | ..................... | .................... | .................... | .................... |
| V | 1474 | hCoV-19/Japan/PG-4936/2020\|EPI_ISL_690056\|2020-07 | .................... | .................... | ..................... | .................... | .................... | .................... |
| V | 1475 | hCoV-19/Japan/PG-4937/2020\|EPI_ISL_690057\|2020-07 | .................... | .................... | ..................... | .................... | .................... | .................... |
| V | 1476 | hCoV-19/Japan/PG-4939/2020\|EPI_ISL_690058\|2020-07 | .................... | .................... | ..................... | .................... | .................... | .................... |
| V | 1477 | hCoV-19/Japan/PG-4940/2020\|EPI_ISL_690059\|2020-07 | .................... | .................... | ..................... | .................... | .................... | .................... |
| V | 1478 | hCoV-19/Japan/PG-4941/2020\|EPI_ISL_690060\|2020-07 | .................... | .................... | ..................... | .................... | .................... | .................... |
| V | 1479 | hCoV-19/Japan/PG-4942/2020\|EPI_ISL_690061\|2020-07 | .................... | .................... | ..................... | .................... | .................... | .................... |
| V | 1480 | hCoV-19/Japan/PG-4943/2020\|EPI_ISL_690062\|2020-07 | .................... | .................... | ..................... | .................... | .................... | .................... |
| V | 1481 | hCoV-19/Japan/PG-4944/2020\|EPI_ISL_690063\|2020-08 | .................... | .................... | ..................... | .................... | .................... | .................... |
| V | 1482 | hCoV-19/Japan/PG-4945/2020\|EPI_ISL_690064\|2020-08 | .................... | .................... | ..................... | .................... | .................... | .................... |
| V | 1483 | hCoV-19/Japan/PG-4946/2020\|EPI_ISL_690065\|2020-08 | .................... | .................... | ..................... | .................... | .................... | .................... |
| V | 1484 | hCoV-19/Japan/PG-4947/2020\|EPI_ISL_690066\|2020-08 | .................... | .................... | ..................... | .................... | .................... | .................... |
| V | 1485 | hCoV-19/Japan/PG-4948/2020\|EPI_ISL_690067\|2020-08 | .................... | .................... | ..................... | .................... | .................... | .................... |
| V | 1486 | hCoV-19/Japan/PG-4949/2020\|EPI_ISL_690068\|2020-08 | .................... | .................... | ..................... | .................... | .................... | .................... |
| V | 1487 | hCoV-19/Japan/PG-4950/2020\|EPI_ISL_690069\|2020-08 | .................... | .................... | ..................... | .................... | .................... | .................... |
| V | 1488 | hCoV-19/Japan/PG-4951/2020\|EPI_ISL_690070\|2020-08 | .................... | .................... | ..................... | .................... | .................... | .................... |
| V | 1489 | hCoV-19/Japan/PG-4952/2020\|EPI_ISL_690071\|2020-08 | .................... | .................... | ..................... | .................... | .................... | .................... |
| V | 1490 | hCoV-19/Japan/PG-4953/2020\|EPI_ISL_690072\|2020-08 | .................... | .................... | ..................... | .................... | .................... | .................... |

| | | | | | | | | |
|---|---|---|---|---|---|---|---|---|
| V | 1491 | hCoV-19/Japan/PG-4954/2020\|EPI_ISL_690073\|2020-08 | .................... | .................... | .................... | .................... | .................... | .................... |
| V | 1492 | hCoV-19/Japan/PG-4955/2020\|EPI_ISL_690074\|2020-08 | .................... | .................... | .................... | .................... | .................... | .................... |
| V | 1493 | hCoV-19/Japan/PG-4956/2020\|EPI_ISL_690075\|2020-08 | .................... | .................... | .................... | .................... | .................... | .................... |
| V | 1494 | hCoV-19/Japan/PG-3248/2020\|EPI_ISL_691363\|2020-04 | .................... | .................... | .................... | .................... | .................... | .................... |
| V | 1495 | hCoV-19/Japan/PG-3249/2020\|EPI_ISL_691364\|2020-06 | .................... | .................... | .................... | .................... | .................... | .................... |
| V | 1496 | hCoV-19/Japan/PG-3250/2020\|EPI_ISL_691365\|2020-06 | .................... | .................... | .................... | .................... | .................... | .................... |
| V | 1497 | hCoV-19/Japan/PG-3251/2020\|EPI_ISL_691366\|2020-06 | .................... | .................... | .................... | .................... | .................... | .................... |
| V | 1498 | hCoV-19/Japan/PG-3252/2020\|EPI_ISL_691367\|2020-06 | .................... | .................... | .................... | .................... | .................... | .................... |
| V | 1499 | hCoV-19/Japan/PG-3253/2020\|EPI_ISL_691368\|2020-06 | .................... | .................... | .................... | .................... | .................... | .................... |
| V | 1500 | hCoV-19/Japan/PG-3254/2020\|EPI_ISL_691369\|2020-06 | .................... | .................... | .................... | .................... | .................... | .................... |
| V | 1501 | hCoV-19/Japan/PG-3255/2020\|EPI_ISL_691370\|2020-06 | .................... | .................... | .................... | .................... | .................... | .................... |
| V | 1502 | hCoV-19/Japan/PG-3256/2020\|EPI_ISL_691371\|2020-06 | .................... | .................... | .................... | .................... | .................... | .................... |
| V | 1503 | hCoV-19/Japan/PG-3257/2020\|EPI_ISL_691372\|2020-06 | .................... | .................... | .................... | .................... | .................... | .................... |
| V | 1504 | hCoV-19/Japan/PG-3258/2020\|EPI_ISL_691373\|2020-06 | .................... | .................... | .................... | .................... | .................... | .................... |
| V | 1505 | hCoV-19/Japan/PG-3259/2020\|EPI_ISL_691374\|2020-06 | .................... | .................... | .................... | .................... | .................... | .................... |
| V | 1506 | hCoV-19/Japan/PG-3260/2020\|EPI_ISL_691375\|2020-06 | .................... | .................... | .................... | .................... | .................... | .................... |
| V | 1507 | hCoV-19/Japan/PG-3261/2020\|EPI_ISL_691376\|2020-06 | .................... | .................... | .................... | .................... | .................... | .................... |
| V | 1508 | hCoV-19/Japan/PG-3262/2020\|EPI_ISL_691377\|2020-06 | .................... | .................... | .................... | .................... | .................... | .................... |
| V | 1509 | hCoV-19/Japan/PG-3263/2020\|EPI_ISL_691378\|2020-07 | .................... | .................... | .................... | .................... | .................... | .................... |
| V | 1510 | hCoV-19/Japan/PG-3264/2020\|EPI_ISL_691379\|2020-03 | .................... | .................... | .................... | .................... | .................... | .................... |
| V | 1511 | hCoV-19/Japan/PG-3265/2020\|EPI_ISL_691380\|2020-03 | .................... | .................... | .................... | .................... | .................... | .................... |
| V | 1512 | hCoV-19/Japan/PG-3266/2020\|EPI_ISL_691381\|2020-03 | .................... | .................... | .................... | .................... | .................... | .................... |
| V | 1513 | hCoV-19/Japan/PG-3267/2020\|EPI_ISL_691382\|2020-03 | .................... | .................... | .................... | .................... | .................... | .................... |
| V | 1514 | hCoV-19/Japan/PG-3268/2020\|EPI_ISL_691383\|2020-03 | .................... | .................... | .................... | .................... | .................... | .................... |
| V | 1515 | hCoV-19/Japan/PG-3269/2020\|EPI_ISL_691384\|2020-04 | .................... | .................... | .................... | .................... | .................... | .................... |
| V | 1516 | hCoV-19/Japan/PG-3270/2020\|EPI_ISL_691385\|2020-04 | .................... | .................... | .................... | .................... | .................... | .................... |
| V | 1517 | hCoV-19/Japan/PG-3271/2020\|EPI_ISL_691386\|2020-07 | .................... | .................... | .................... | .................... | .................... | .................... |
| V | 1518 | hCoV-19/Japan/PG-3272/2020\|EPI_ISL_691387\|2020-07 | .................... | .................... | .................... | .................... | .................... | .................... |
| V | 1519 | hCoV-19/Japan/PG-3273/2020\|EPI_ISL_691388\|2020-07 | .................... | .................... | .................... | .................... | .................... | .................... |
| V | 1520 | hCoV-19/Japan/PG-3274/2020\|EPI_ISL_691389\|2020-07 | .................... | .................... | .................... | .................... | .................... | .................... |
| V | 1521 | hCoV-19/Japan/PG-3275/2020\|EPI_ISL_691390\|2020-07 | .................... | .................... | .................... | .................... | .................... | .................... |
| V | 1522 | hCoV-19/Japan/PG-3276/2020\|EPI_ISL_691391\|2020-07 | .................... | .................... | .................... | .................... | .................... | .................... |
| V | 1523 | hCoV-19/Japan/PG-3277/2020\|EPI_ISL_691392\|2020-07 | .................... | .................... | .................... | .................... | .................... | .................... |
| V | 1524 | hCoV-19/Japan/PG-3278/2020\|EPI_ISL_691393\|2020-07 | .................... | .................... | .................... | .................... | .................... | .................... |
| V | 1525 | hCoV-19/Japan/PG-3279/2020\|EPI_ISL_691394\|2020-07 | .................... | .................... | .................... | .................... | .................... | .................... |
| V | 1526 | hCoV-19/Japan/PG-3280/2020\|EPI_ISL_691395\|2020-03 | .................... | .................... | .................... | .................... | .................... | .................... |
| V | 1527 | hCoV-19/Japan/PG-3281/2020\|EPI_ISL_691396\|2020-04 | .................... | .................... | .................... | .................... | .................... | .................... |
| V | 1528 | hCoV-19/Japan/PG-3282/2020\|EPI_ISL_691397\|2020-04 | .................... | .................... | .................... | .................... | .................... | .................... |
| V | 1529 | hCoV-19/Japan/PG-3283/2020\|EPI_ISL_691398\|2020-04 | .................... | .................... | .................... | .................... | .................... | .................... |
| V | 1530 | hCoV-19/Japan/PG-3284/2020\|EPI_ISL_691399\|2020-04 | .................... | .................... | .................... | .................... | .................... | .................... |
| V | 1531 | hCoV-19/Japan/PG-4471/2020\|EPI_ISL_692694\|2020-07 | .................... | .................... | .................... | .................... | .................... | .................... |
| V | 1532 | hCoV-19/Japan/PG-4472/2020\|EPI_ISL_692695\|2020-07 | .................... | .................... | .................... | .................... | .................... | .................... |
| V | 1533 | hCoV-19/Japan/PG-4473/2020\|EPI_ISL_692696\|2020-07 | .................... | .................... | .................... | .................... | .................... | .................... |
| V | 1534 | hCoV-19/Japan/PG-4474/2020\|EPI_ISL_692697\|2020-07 | .................... | .................... | .................... | .................... | .................... | .................... |
| V | 1535 | hCoV-19/Japan/PG-4475/2020\|EPI_ISL_692698\|2020-07 | .................... | .................... | .................... | .................... | .................... | .................... |
| V | 1537 | hCoV-19/Japan/PG-0586/2020\|EPI_ISL_684200\|2020-04 | .................... | .................... | .................... | .................... | .................... | .................... |
| V | 1538 | hCoV-19/Japan/PG-0588/2020\|EPI_ISL_684201\|2020-04 | .................... | .................... | .................... | .................... | .................... | .................... |
| V | 1539 | hCoV-19/Japan/PG-0592/2020\|EPI_ISL_684202\|2020-04 | .................... | .................... | .................... | .................... | .................... | .................... |
| V | 1540 | hCoV-19/Japan/PG-0593/2020\|EPI_ISL_684203\|2020-04 | .................... | .................... | .................... | .................... | .................... | .................... |
| V | 1541 | hCoV-19/Japan/PG-0594/2020\|EPI_ISL_684204\|2020-04 | .................... | .................... | .................... | .................... | .................... | .................... |
| V | 1542 | hCoV-19/Japan/PG-0595/2020\|EPI_ISL_684205\|2020-04 | .................... | .................... | .................... | .................... | .................... | .................... |
| V | 1543 | hCoV-19/Japan/PG-0596/2020\|EPI_ISL_684206\|2020-04 | .................... | .................... | .................... | .................... | .................... | .................... |
| V | 1544 | hCoV-19/Japan/PG-0597/2020\|EPI_ISL_684207\|2020-04 | .................... | .................... | .................... | .................... | .................... | .................... |
| V | 1545 | hCoV-19/Japan/PG-0598/2020\|EPI_ISL_684208\|2020-04 | .................... | .................... | .................... | .................... | .................... | .................... |
| V | 1546 | hCoV-19/Japan/PG-0599/2020\|EPI_ISL_684209\|2020-04 | .................... | .................... | .................... | .................... | .................... | .................... |
| V | 1547 | hCoV-19/Japan/PG-0600/2020\|EPI_ISL_684210\|2020-04 | .................... | .................... | .................... | .................... | .................... | .................... |
| V | 1548 | hCoV-19/Japan/PG-0601/2020\|EPI_ISL_684211\|2020-04 | .................... | .................... | .................... | .................... | .................... | .................... |
| V | 1549 | hCoV-19/Japan/PG-0602/2020\|EPI_ISL_684212\|2020-04 | .................... | .................... | .................... | .................... | .................... | .................... |
| V | 1550 | hCoV-19/Japan/PG-0603/2020\|EPI_ISL_684213\|2020-04 | .................... | .................... | .................... | .................... | .................... | .................... |
| V | 1551 | hCoV-19/Japan/PG-0604/2020\|EPI_ISL_684214\|2020-04 | .................... | .................... | .................... | .................... | .................... | .................... |
| V | 1552 | hCoV-19/Japan/PG-0605/2020\|EPI_ISL_684215\|2020-04 | .................... | .................... | .................... | .................... | .................... | .................... |
| V | 1553 | hCoV-19/Japan/PG-0606/2020\|EPI_ISL_684216\|2020-04 | .................... | .................... | .................... | .................... | .................... | .................... |
| V | 1554 | hCoV-19/Japan/PG-0607/2020\|EPI_ISL_684217\|2020-04 | .................... | .................... | .................... | .................... | .................... | .................... |
| V | 1555 | hCoV-19/Japan/PG-0608/2020\|EPI_ISL_684218\|2020-04 | .................... | .................... | .................... | .................... | .................... | .................... |
| V | 1556 | hCoV-19/Japan/PG-0609/2020\|EPI_ISL_684219\|2020-03 | .................... | .................... | .................... | .................... | .................... | .................... |
| V | 1557 | hCoV-19/Japan/PG-0610/2020\|EPI_ISL_684220\|2020-04 | .................... | .................... | .................... | .................... | .................... | .................... |
| V | 1558 | hCoV-19/Japan/PG-0611/2020\|EPI_ISL_684221\|2020-04 | .................... | .................... | .................... | .................... | .................... | .................... |
| V | 1559 | hCoV-19/Japan/PG-0612/2020\|EPI_ISL_684222\|2020-04 | .................... | .................... | .................... | .................... | .................... | .................... |
| V | 1560 | hCoV-19/Japan/PG-0613/2020\|EPI_ISL_684223\|2020-04 | .................... | .................... | .................... | .................... | .................... | .................... |
| V | 1561 | hCoV-19/Japan/PG-0614/2020\|EPI_ISL_684224\|2020-04 | .................... | .................... | .................... | .................... | .................... | .................... |
| V | 1562 | hCoV-19/Japan/PG-0615/2020\|EPI_ISL_684225\|2020-04 | .................... | .................... | .................... | .................... | .................... | .................... |
| V | 1563 | hCoV-19/Japan/PG-0616/2020\|EPI_ISL_684226\|2020-04 | .................... | .................... | .................... | .................... | .................... | .................... |
| V | 1564 | hCoV-19/Japan/PG-0617/2020\|EPI_ISL_684227\|2020-04 | .................... | .................... | .................... | .................... | .................... | .................... |
| V | 1565 | hCoV-19/Japan/PG-0618/2020\|EPI_ISL_684228\|2020-04 | .................... | .................... | .................... | .................... | .................... | .................... |
| V | 1566 | hCoV-19/Japan/PG-0619/2020\|EPI_ISL_684229\|2020-04 | .................... | .................... | .................... | .................... | .................... | .................... |
| V | 1567 | hCoV-19/Japan/PG-0620/2020\|EPI_ISL_684230\|2020-04 | .................... | .................... | .................... | .................... | .................... | .................... |
| V | 1568 | hCoV-19/Japan/PG-0621/2020\|EPI_ISL_684231\|2020-04 | .................... | .................... | .................... | .................... | .................... | .................... |
| V | 1569 | hCoV-19/Japan/PG-0622/2020\|EPI_ISL_684232\|2020-04 | .................... | .................... | .................... | .................... | .................... | .................... |
| V | 1570 | hCoV-19/Japan/PG-0623/2020\|EPI_ISL_684233\|2020-04 | .................... | .................... | .................... | .................... | .................... | .................... |
| V | 1571 | hCoV-19/Japan/PG-0624/2020\|EPI_ISL_684234\|2020-04 | .................... | .................... | .................... | .................... | .................... | .................... |

| | | | | | | | | |
|---|---|---|---|---|---|---|---|---|
| V | 1572 | hCoV-19/Japan/PG-0625/2020\|EPI_ISL_684235\|2020-04 | .................... | .................... | ..................... | .................... | .................... | .................... |
| V | 1573 | hCoV-19/Japan/PG-0626/2020\|EPI_ISL_684236\|2020-04 | .................... | .................... | ..................... | .................... | .................... | .................... |
| V | 1574 | hCoV-19/Japan/PG-0627/2020\|EPI_ISL_684237\|2020-04 | .................... | .................... | ..................... | .................... | .................... | .................... |
| V | 1575 | hCoV-19/Japan/PG-0628/2020\|EPI_ISL_684238\|2020-04 | .................... | .................... | ..................... | .................... | .................... | .................... |
| V | 1576 | hCoV-19/Japan/PG-1916/2020\|EPI_ISL_685522\|2020-05 | .................... | .................... | ..................... | .................... | .................... | .................... |
| V | 1577 | hCoV-19/Japan/PG-1917/2020\|EPI_ISL_685523\|2020-05 | .................... | .................... | ..................... | .................... | .................... | .................... |
| V | 1578 | hCoV-19/Japan/PG-1918/2020\|EPI_ISL_685524\|2020-03 | .................... | .................... | ..................... | .................... | .................... | .................... |
| V | 1579 | hCoV-19/Japan/PG-1919/2020\|EPI_ISL_685525\|2020-03 | .................... | .................... | ..................... | .................... | .................... | .................... |
| V | 1580 | hCoV-19/Japan/PG-1920/2020\|EPI_ISL_685526\|2020-03 | .................... | .................... | ..................... | .................... | .................... | .................... |
| V | 1581 | hCoV-19/Japan/PG-1921/2020\|EPI_ISL_685527\|2020-03 | .................... | .................... | ..................... | .................... | .................... | .................... |
| V | 1582 | hCoV-19/Japan/PG-1922/2020\|EPI_ISL_685528\|2020-04 | .................... | .................... | ..................... | .................... | .................... | .................... |
| V | 1583 | hCoV-19/Japan/PG-1923/2020\|EPI_ISL_685529\|2020-04 | .................... | .................... | ..................... | .................... | .................... | .................... |
| V | 1584 | hCoV-19/Japan/PG-1924/2020\|EPI_ISL_685530\|2020-04 | .................... | .................... | ..................... | .................... | .................... | .................... |
| V | 1585 | hCoV-19/Japan/PG-1925/2020\|EPI_ISL_685531\|2020-04 | .................... | .................... | ..................... | .................... | .................... | .................... |
| V | 1586 | hCoV-19/Japan/PG-1926/2020\|EPI_ISL_685532\|2020-04 | .................... | .................... | ..................... | .................... | .................... | .................... |
| V | 1587 | hCoV-19/Japan/PG-1927/2020\|EPI_ISL_685533\|2020-04 | .................... | .................... | ..................... | .................... | .................... | .................... |
| V | 1588 | hCoV-19/Japan/PG-1928/2020\|EPI_ISL_685534\|2020-04 | .................... | .................... | ..................... | .................... | .................... | .................... |
| V | 1589 | hCoV-19/Japan/PG-1929/2020\|EPI_ISL_685535\|2020-04 | .................... | .................... | ..................... | .................... | .................... | .................... |
| V | 1590 | hCoV-19/Japan/PG-1930/2020\|EPI_ISL_685536\|2020-04 | .................... | .................... | ..................... | .................... | .................... | .................... |
| V | 1591 | hCoV-19/Japan/PG-1931/2020\|EPI_ISL_685537\|2020-04 | .................... | .................... | ..................... | .................... | .................... | .................... |
| V | 1592 | hCoV-19/Japan/PG-1932/2020\|EPI_ISL_685538\|2020-04 | .................... | .................... | ..................... | .................... | .................... | .................... |
| V | 1593 | hCoV-19/Japan/PG-1933/2020\|EPI_ISL_685539\|2020-04 | .................... | .................... | ..................... | .................... | .................... | .................... |
| V | 1594 | hCoV-19/Japan/PG-1934/2020\|EPI_ISL_685540\|2020-04 | .................... | .................... | ..................... | .................... | .................... | .................... |
| V | 1595 | hCoV-19/Japan/PG-1935/2020\|EPI_ISL_685541\|2020-04 | .................... | .................... | ..................... | .................... | .................... | .................... |
| V | 1596 | hCoV-19/Japan/PG-1936/2020\|EPI_ISL_685542\|2020-04 | .................... | .................... | ..................... | .................... | .................... | .................... |
| V | 1597 | hCoV-19/Japan/PG-1937/2020\|EPI_ISL_685543\|2020-04 | .................... | .................... | ..................... | .................... | .................... | .................... |
| V | 1598 | hCoV-19/Japan/PG-1938/2020\|EPI_ISL_685544\|2020-04 | .................... | .................... | ..................... | .................... | .................... | .................... |
| V | 1599 | hCoV-19/Japan/PG-1939/2020\|EPI_ISL_685545\|2020-04 | .................... | .................... | ..................... | .................... | .................... | .................... |
| V | 1600 | hCoV-19/Japan/PG-1940/2020\|EPI_ISL_685546\|2020-04 | .................... | .................... | ..................... | .................... | .................... | .................... |
| V | 1601 | hCoV-19/Japan/PG-1941/2020\|EPI_ISL_685547\|2020-04 | .................... | .................... | ..................... | .................... | .................... | .................... |
| V | 1602 | hCoV-19/Japan/PG-1942/2020\|EPI_ISL_685548\|2020-04 | .................... | .................... | ..................... | .................... | .................... | .................... |
| V | 1603 | hCoV-19/Japan/PG-1943/2020\|EPI_ISL_685549\|2020-04 | .................... | .................... | ..................... | .................... | .................... | .................... |
| V | 1604 | hCoV-19/Japan/PG-1944/2020\|EPI_ISL_685550\|2020-04 | .................... | .................... | ..................... | .................... | .................... | .................... |
| V | 1605 | hCoV-19/Japan/PG-1945/2020\|EPI_ISL_685551\|2020-04 | .................... | .................... | ..................... | .................... | .................... | .................... |
| V | 1606 | hCoV-19/Japan/PG-1946/2020\|EPI_ISL_685552\|2020-04 | .................... | .................... | ..................... | .................... | .................... | .................... |
| V | 1607 | hCoV-19/Japan/PG-1947/2020\|EPI_ISL_685553\|2020-04 | .................... | .................... | ..................... | .................... | .................... | .................... |
| V | 1608 | hCoV-19/Japan/PG-1948/2020\|EPI_ISL_685554\|2020-04 | .................... | .................... | ..................... | .................... | .................... | .................... |
| V | 1609 | hCoV-19/Japan/PG-1949/2020\|EPI_ISL_685555\|2020-04 | .................... | .................... | ..................... | .................... | .................... | .................... |
| V | 1610 | hCoV-19/Japan/PG-1950/2020\|EPI_ISL_685556\|2020-04 | .................... | .................... | ..................... | .................... | .................... | .................... |
| V | 1611 | hCoV-19/Japan/PG-1951/2020\|EPI_ISL_685557\|2020-04 | .................... | .................... | ..................... | .................... | .................... | .................... |
| V | 1612 | hCoV-19/Japan/PG-1952/2020\|EPI_ISL_685558\|2020-04 | .................... | .................... | ..................... | .................... | .................... | .................... |
| V | 1613 | hCoV-19/Japan/PG-1953/2020\|EPI_ISL_685559\|2020-04 | .................... | .................... | ..................... | .................... | .................... | .................... |
| V | 1614 | hCoV-19/Japan/PG-1954/2020\|EPI_ISL_685560\|2020-04 | .................... | .................... | ..................... | .................... | .................... | .................... |
| V | 1615 | hCoV-19/Japan/PG-1955/2020\|EPI_ISL_685561\|2020-04 | .................... | .................... | ..................... | .................... | .................... | .................... |
| V | 1616 | hCoV-19/Japan/PG-1956/2020\|EPI_ISL_685562\|2020-04 | .................... | .................... | ..................... | .................... | .................... | .................... |
| V | 1617 | hCoV-19/Japan/PG-1957/2020\|EPI_ISL_685563\|2020-04 | .................... | .................... | ..................... | .................... | .................... | .................... |
| V | 1618 | hCoV-19/Japan/PG-1958/2020\|EPI_ISL_685564\|2020-04 | .................... | .................... | ..................... | .................... | .................... | .................... |
| V | 1619 | hCoV-19/Japan/PG-1959/2020\|EPI_ISL_685565\|2020-04 | .................... | .................... | ..................... | .................... | .................... | .................... |
| V | 1620 | hCoV-19/Japan/PG-1960/2020\|EPI_ISL_685566\|2020-04 | .................... | .................... | ..................... | .................... | .................... | .................... |
| V | 1621 | hCoV-19/Japan/PG-1961/2020\|EPI_ISL_685567\|2020-04 | .................... | .................... | ..................... | .................... | .................... | .................... |
| V | 1622 | hCoV-19/Japan/PG-1962/2020\|EPI_ISL_685568\|2020-04 | .................... | .................... | ..................... | .................... | .................... | .................... |
| V | 1623 | hCoV-19/Japan/PG-1963/2020\|EPI_ISL_685569\|2020-04 | .................... | .................... | ..................... | .................... | .................... | .................... |
| V | 1624 | hCoV-19/Japan/PG-7751/2020\|EPI_ISL_686853\|2020-04 | .................... | .................... | ..................... | .................... | .................... | .................... |
| V | 1625 | hCoV-19/Japan/PG-7752/2020\|EPI_ISL_686854\|2020-04 | .................... | .................... | ..................... | .................... | .................... | .................... |
| V | 1626 | hCoV-19/Japan/PG-7753/2020\|EPI_ISL_686855\|2020-04 | .................... | .................... | ..................... | .................... | .................... | .................... |
| V | 1627 | hCoV-19/Japan/PG-7754/2020\|EPI_ISL_686856\|2020-04 | .................... | .................... | ..................... | .................... | .................... | .................... |
| V | 1628 | hCoV-19/Japan/PG-7755/2020\|EPI_ISL_686857\|2020-04 | .................... | .................... | ..................... | .................... | .................... | .................... |
| V | 1629 | hCoV-19/Japan/PG-7756/2020\|EPI_ISL_686858\|2020-04 | .................... | .................... | ..................... | .................... | .................... | .................... |
| V | 1630 | hCoV-19/Japan/PG-7757/2020\|EPI_ISL_686859\|2020-04 | .................... | .................... | ..................... | .................... | .................... | .................... |
| V | 1631 | hCoV-19/Japan/PG-7758/2020\|EPI_ISL_686860\|2020-04 | .................... | .................... | ..................... | .................... | .................... | .................... |
| V | 1632 | hCoV-19/Japan/PG-7759/2020\|EPI_ISL_686861\|2020-04 | .................... | .................... | ..................... | .................... | .................... | .................... |
| V | 1633 | hCoV-19/Japan/PG-7760/2020\|EPI_ISL_686862\|2020-04 | .................... | .................... | ..................... | .................... | .................... | .................... |
| V | 1634 | hCoV-19/Japan/PG-7761/2020\|EPI_ISL_686863\|2020-04 | .................... | .................... | ..................... | .................... | .................... | .................... |
| V | 1635 | hCoV-19/Japan/PG-7762/2020\|EPI_ISL_686864\|2020-04 | .................... | .................... | ..................... | .................... | .................... | .................... |
| V | 1636 | hCoV-19/Japan/PG-7763/2020\|EPI_ISL_686865\|2020-04 | .................... | .................... | ..................... | .................... | .................... | .................... |
| V | 1637 | hCoV-19/Japan/PG-7764/2020\|EPI_ISL_686866\|2020-04 | .................... | .................... | ..................... | .................... | .................... | .................... |
| V | 1638 | hCoV-19/Japan/PG-7765/2020\|EPI_ISL_686867\|2020-04 | .................... | .................... | ..................... | .................... | .................... | .................... |
| V | 1639 | hCoV-19/Japan/PG-7766/2020\|EPI_ISL_686868\|2020-04 | .................... | .................... | ..................... | .................... | .................... | .................... |
| V | 1640 | hCoV-19/Japan/PG-7767/2020\|EPI_ISL_686869\|2020-04 | .................... | .................... | ..................... | .................... | .................... | .................... |
| V | 1641 | hCoV-19/Japan/PG-7768/2020\|EPI_ISL_686870\|2020-04 | .................... | .................... | ..................... | .................... | .................... | .................... |
| V | 1642 | hCoV-19/Japan/PG-7769/2020\|EPI_ISL_686871\|2020-04 | .................... | .................... | ..................... | .................... | .................... | .................... |
| V | 1643 | hCoV-19/Japan/PG-7770/2020\|EPI_ISL_686872\|2020-04 | .................... | .................... | ..................... | .................... | .................... | .................... |
| V | 1644 | hCoV-19/Japan/PG-7771/2020\|EPI_ISL_686873\|2020-04 | .................... | .................... | ..................... | .................... | .................... | .................... |
| V | 1645 | hCoV-19/Japan/PG-7772/2020\|EPI_ISL_686874\|2020-06 | .................... | .................... | ..................... | .................... | .................... | .................... |
| V | 1646 | hCoV-19/Japan/PG-7773/2020\|EPI_ISL_686875\|2020-06 | .................... | .................... | ..................... | .................... | .................... | .................... |
| V | 1647 | hCoV-19/Japan/PG-7774/2020\|EPI_ISL_686876\|2020-06 | .................... | .................... | ..................... | .................... | .................... | .................... |
| V | 1648 | hCoV-19/Japan/PG-7775/2020\|EPI_ISL_686877\|2020-07 | .................... | .................... | ..................... | .................... | .................... | .................... |
| V | 1649 | hCoV-19/Japan/PG-7776/2020\|EPI_ISL_686878\|2020-07 | .................... | .................... | ..................... | .................... | .................... | .................... |
| V | 1650 | hCoV-19/Japan/PG-7777/2020\|EPI_ISL_686879\|2020-07 | .................... | .................... | ..................... | .................... | .................... | .................... |
| V | 1651 | hCoV-19/Japan/PG-7778/2020\|EPI_ISL_686880\|2020-07 | .................... | .................... | ..................... | .................... | .................... | .................... |

| | | | | | | | |
|---|---|---|---|---|---|---|---|
| V | 1652 hCoV-19/Japan/PG-7779/2020\|EPI_ISL_686881\|2020-07 | .................... | .................... | ..................... | .................... | .................... | .................... |
| V | 1653 hCoV-19/Japan/PG-7780/2020\|EPI_ISL_686882\|2020-07 | .................... | .................... | ..................... | .................... | .................... | .................... |
| V | 1654 hCoV-19/Japan/PG-7781/2020\|EPI_ISL_686883\|2020-07 | .................... | .................... | ..................... | .................... | .................... | .................... |
| V | 1655 hCoV-19/Japan/PG-7782/2020\|EPI_ISL_686884\|2020-07 | .................... | .................... | ..................... | .................... | .................... | .................... |
| V | 1656 hCoV-19/Japan/PG-7783/2020\|EPI_ISL_686885\|2020-07 | .................... | .................... | ..................... | .................... | .................... | .................... |
| V | 1657 hCoV-19/Japan/PG-7784/2020\|EPI_ISL_686886\|2020-07 | .................... | .................... | ..................... | .................... | .................... | .................... |
| V | 1658 hCoV-19/Japan/PG-7785/2020\|EPI_ISL_686887\|2020-07 | .................... | .................... | ..................... | .................... | .................... | .................... |
| V | 1659 hCoV-19/Japan/PG-7786/2020\|EPI_ISL_686888\|2020-07 | .................... | .................... | ..................... | .................... | .................... | .................... |
| V | 1660 hCoV-19/Japan/PG-7787/2020\|EPI_ISL_686889\|2020-07 | .................... | .................... | ..................... | .................... | .................... | .................... |
| V | 1661 hCoV-19/Japan/PG-7788/2020\|EPI_ISL_686890\|2020-07 | .................... | .................... | ..................... | .................... | .................... | .................... |
| V | 1662 hCoV-19/Japan/PG-7789/2020\|EPI_ISL_686891\|2020-07 | .................... | .................... | ..................... | .................... | .................... | .................... |
| V | 1663 hCoV-19/Japan/PG-7790/2020\|EPI_ISL_686892\|2020-07 | .................... | .................... | ..................... | .................... | .................... | .................... |
| V | 1664 hCoV-19/Japan/PG-7791/2020\|EPI_ISL_686893\|2020-07 | .................... | .................... | ..................... | .................... | .................... | .................... |
| V | 1665 hCoV-19/Japan/PG-7792/2020\|EPI_ISL_686894\|2020-07 | .................... | .................... | ..................... | .................... | .................... | .................... |
| V | 1666 hCoV-19/Japan/PG-7793/2020\|EPI_ISL_686895\|2020-07 | .................... | .................... | ..................... | .................... | .................... | .................... |
| V | 1667 hCoV-19/Japan/PG-7794/2020\|EPI_ISL_686896\|2020-07 | .................... | .................... | ..................... | .................... | .................... | .................... |
| V | 1668 hCoV-19/Japan/PG-7795/2020\|EPI_ISL_686897\|2020-07 | .................... | .................... | ..................... | .................... | .................... | .................... |
| V | 1669 hCoV-19/Japan/PG-7796/2020\|EPI_ISL_686898\|2020-07 | .................... | .................... | ..................... | .................... | .................... | .................... |
| V | 1670 hCoV-19/Japan/PG-7797/2020\|EPI_ISL_686899\|2020-07 | .................... | .................... | ..................... | .................... | .................... | .................... |
| V | 1671 hCoV-19/Japan/PG-4957/2020\|EPI_ISL_690076\|2020-08 | .................... | .................... | ..................... | .................... | .................... | .................... |
| V | 1672 hCoV-19/Japan/PG-4958/2020\|EPI_ISL_690077\|2020-08 | .................... | .................... | ..................... | .................... | .................... | .................... |
| V | 1673 hCoV-19/Japan/PG-4959/2020\|EPI_ISL_690078\|2020-08 | .................... | .................... | ..................... | .................... | .................... | .................... |
| V | 1674 hCoV-19/Japan/PG-4960/2020\|EPI_ISL_690079\|2020-08 | .................... | .................... | ..................... | .................... | .................... | .................... |
| V | 1675 hCoV-19/Japan/PG-4962/2020\|EPI_ISL_690080\|2020-07 | .................... | .................... | ..................... | .................... | .................... | .................... |
| V | 1676 hCoV-19/Japan/PG-4963/2020\|EPI_ISL_690081\|2020-07 | .................... | .................... | ..................... | .................... | .................... | .................... |
| V | 1677 hCoV-19/Japan/PG-4964/2020\|EPI_ISL_690082\|2020-07 | .................... | .................... | ..................... | .................... | .................... | .................... |
| V | 1678 hCoV-19/Japan/PG-4965/2020\|EPI_ISL_690083\|2020-07 | .................... | .................... | ..................... | .................... | .................... | .................... |
| V | 1679 hCoV-19/Japan/PG-4966/2020\|EPI_ISL_690084\|2020-07 | .................... | .................... | ..................... | .................... | .................... | .................... |
| V | 1680 hCoV-19/Japan/PG-4967/2020\|EPI_ISL_690085\|2020-08 | .................... | .................... | ..................... | .................... | .................... | .................... |
| V | 1681 hCoV-19/Japan/PG-4968/2020\|EPI_ISL_690086\|2020-08 | .................... | .................... | ..................... | .................... | .................... | .................... |
| V | 1682 hCoV-19/Japan/PG-4969/2020\|EPI_ISL_690087\|2020-08 | .................... | .................... | ..................... | .................... | .................... | .................... |
| V | 1683 hCoV-19/Japan/PG-4970/2020\|EPI_ISL_690088\|2020-08 | .................... | .................... | ..................... | .................... | .................... | .................... |
| V | 1684 hCoV-19/Japan/PG-4971/2020\|EPI_ISL_690089\|2020-08 | .................... | .................... | ..................... | .................... | .................... | .................... |
| V | 1685 hCoV-19/Japan/PG-4972/2020\|EPI_ISL_690090\|2020-08 | .................... | .................... | ..................... | .................... | .................... | .................... |
| V | 1686 hCoV-19/Japan/PG-4973/2020\|EPI_ISL_690091\|2020-08 | .................... | .................... | ..................... | .................... | .................... | .................... |
| V | 1687 hCoV-19/Japan/PG-4974/2020\|EPI_ISL_690092\|2020-08 | .................... | .................... | ..................... | .................... | .................... | .................... |
| V | 1688 hCoV-19/Japan/PG-4976/2020\|EPI_ISL_690093\|2020-08 | .................... | .................... | ..................... | .................... | .................... | .................... |
| V | 1689 hCoV-19/Japan/PG-4978/2020\|EPI_ISL_690094\|2020-08 | .................... | .................... | ..................... | .................... | .................... | .................... |
| V | 1690 hCoV-19/Japan/PG-4979/2020\|EPI_ISL_690095\|2020-08 | .................... | .................... | ..................... | .................... | .................... | .................... |
| V | 1691 hCoV-19/Japan/PG-4980/2020\|EPI_ISL_690096\|2020-08 | .................... | .................... | ..................... | .................... | .................... | .................... |
| V | 1692 hCoV-19/Japan/PG-4981/2020\|EPI_ISL_690097\|2020-08 | .................... | .................... | ..................... | .................... | .................... | .................... |
| V | 1693 hCoV-19/Japan/PG-4984/2020\|EPI_ISL_690098\|2020-08 | .................... | .................... | ..................... | .................... | .................... | .................... |
| V | 1694 hCoV-19/Japan/PG-4986/2020\|EPI_ISL_690099\|2020-08 | .................... | .................... | ..................... | .................... | .................... | .................... |
| V | 1695 hCoV-19/Japan/PG-3180/2020\|EPI_ISL_691300\|2020-04 | .................... | .................... | ..................... | .................... | .................... | .................... |
| V | 1696 hCoV-19/Japan/PG-3181/2020\|EPI_ISL_691301\|2020-04 | .................... | .................... | ..................... | .................... | .................... | .................... |
| V | 1697 hCoV-19/Japan/PG-3182/2020\|EPI_ISL_691302\|2020-04 | .................... | .................... | ..................... | .................... | .................... | .................... |
| V | 1698 hCoV-19/Japan/PG-3183/2020\|EPI_ISL_691303\|2020-04 | .................... | .................... | ..................... | .................... | .................... | .................... |
| V | 1699 hCoV-19/Japan/PG-3184/2020\|EPI_ISL_691304\|2020-04 | .................... | .................... | ..................... | .................... | .................... | .................... |
| V | 1700 hCoV-19/Japan/PG-3185/2020\|EPI_ISL_691305\|2020-04 | .................... | .................... | ..................... | .................... | .................... | .................... |
| V | 1701 hCoV-19/Japan/PG-3186/2020\|EPI_ISL_691306\|2020-04 | .................... | .................... | ..................... | .................... | .................... | .................... |
| V | 1702 hCoV-19/Japan/PG-3187/2020\|EPI_ISL_691307\|2020-04 | .................... | .................... | ..................... | .................... | .................... | .................... |
| V | 1703 hCoV-19/Japan/PG-3188/2020\|EPI_ISL_691308\|2020-04 | .................... | .................... | ..................... | .................... | .................... | .................... |
| V | 1704 hCoV-19/Japan/PG-3189/2020\|EPI_ISL_691309\|2020-04 | .................... | .................... | ..................... | .................... | .................... | .................... |
| V | 1705 hCoV-19/Japan/PG-3190/2020\|EPI_ISL_691310\|2020-04 | .................... | .................... | ..................... | .................... | .................... | .................... |
| V | 1706 hCoV-19/Japan/PG-3191/2020\|EPI_ISL_691311\|2020-04 | .................... | .................... | ..................... | .................... | .................... | .................... |
| V | 1707 hCoV-19/Japan/PG-3192/2020\|EPI_ISL_691312\|2020-04 | .................... | .................... | ..................... | .................... | .................... | .................... |
| V | 1708 hCoV-19/Japan/PG-3193/2020\|EPI_ISL_691313\|2020-04 | .................... | .................... | ..................... | .................... | .................... | .................... |
| V | 1709 hCoV-19/Japan/PG-3194/2020\|EPI_ISL_691314\|2020-04 | .................... | .................... | ..................... | .................... | .................... | .................... |
| V | 1710 hCoV-19/Japan/PG-3195/2020\|EPI_ISL_691315\|2020-04 | .................... | .................... | ..................... | .................... | .................... | .................... |
| V | 1711 hCoV-19/Japan/PG-3196/2020\|EPI_ISL_691316\|2020-04 | .................... | .................... | ..................... | .................... | .................... | .................... |
| V | 1712 hCoV-19/Japan/PG-3197/2020\|EPI_ISL_691317\|2020-04 | .................... | .................... | ..................... | .................... | .................... | .................... |
| V | 1713 hCoV-19/Japan/PG-3198/2020\|EPI_ISL_691318\|2020-04 | .................... | .................... | ..................... | .................... | .................... | .................... |
| V | 1714 hCoV-19/Japan/PG-3199/2020\|EPI_ISL_691319\|2020-04 | .................... | .................... | ..................... | .................... | .................... | .................... |
| V | 1715 hCoV-19/Japan/PG-4383/2020\|EPI_ISL_692610\|2020-06 | .................... | .................... | ..................... | .................... | .................... | .................... |
| V | 1716 hCoV-19/Japan/PG-4384/2020\|EPI_ISL_692611\|2020-06 | .................... | .................... | ..................... | .................... | .................... | .................... |
| V | 1717 hCoV-19/Japan/PG-4385/2020\|EPI_ISL_692612\|2020-07 | .................... | .................... | ..................... | .................... | .................... | .................... |
| V | 1718 hCoV-19/Japan/PG-4386/2020\|EPI_ISL_692613\|2020-07 | .................... | .................... | ..................... | .................... | .................... | .................... |
| V | 1719 hCoV-19/Japan/PG-4387/2020\|EPI_ISL_692614\|2020-07 | .................... | .................... | ..................... | .................... | .................... | .................... |
| V | 1721 hCoV-19/Japan/PG-4389/2020\|EPI_ISL_692616\|2020-07 | .................... | .................... | ..................... | .................... | .................... | .................... |
| V | 1722 hCoV-19/Japan/PG-4390/2020\|EPI_ISL_692617\|2020-07 | .................... | .................... | ..................... | .................... | .................... | .................... |
| V | 1723 hCoV-19/Japan/PG-4391/2020\|EPI_ISL_692618\|2020-07 | .................... | .................... | ..................... | .................... | .................... | .................... |
| V | 1724 hCoV-19/Japan/PG-4392/2020\|EPI_ISL_692619\|2020-07 | .................... | .................... | ..................... | .................... | .................... | .................... |
| V | 1725 hCoV-19/Japan/PG-4394/2020\|EPI_ISL_692620\|2020-07 | .................... | .................... | ..................... | .................... | .................... | .................... |
| V | 1726 hCoV-19/Japan/PG-4395/2020\|EPI_ISL_692621\|2020-07 | .................... | .................... | ..................... | .................... | .................... | .................... |
| V | 1727 hCoV-19/Japan/PG-4396/2020\|EPI_ISL_692622\|2020-07 | .................... | .................... | ..................... | .................... | .................... | .................... |
| V | 1728 hCoV-19/Japan/PG-4397/2020\|EPI_ISL_692623\|2020-07 | .................... | .................... | ..................... | .................... | .................... | .................... |
| V | 1729 hCoV-19/Japan/PG-4398/2020\|EPI_ISL_692624\|2020-07 | .................... | .................... | ..................... | .................... | .................... | .................... |
| V | 1731 hCoV-19/Japan/PG-4400/2020\|EPI_ISL_692626\|2020-07 | .................... | .................... | ..................... | .................... | .................... | .................... |
| V | 1732 hCoV-19/Japan/PG-4401/2020\|EPI_ISL_692627\|2020-07 | .................... | .................... | ..................... | .................... | .................... | .................... |
| V | 1733 hCoV-19/Japan/PG-4402/2020\|EPI_ISL_692628\|2020-07 | .................... | .................... | ..................... | .................... | .................... | .................... |

| | | | | | | | | |
|---|---|---|---|---|---|---|---|---|
| V | 1734 | hCoV-19/Japan/PG-4403/2020\|EPI_ISL_692629\|2020-07 | .................... | .................... | ..................... | .................... | .................... | .................... |
| V | 1735 | hCoV-19/Japan/PG-4404/2020\|EPI_ISL_692630\|2020-07 | .................... | .................... | ..................... | .................... | .................... | .................... |
| V | 1737 | hCoV-19/Japan/PG-4406/2020\|EPI_ISL_692632\|2020-07 | .................... | .................... | ..................... | .................... | .................... | .................... |
| V | 1738 | hCoV-19/Japan/PG-4407/2020\|EPI_ISL_692633\|2020-07 | .................... | .................... | ..................... | .................... | .................... | .................... |
| V | 1739 | hCoV-19/Japan/PG-4408/2020\|EPI_ISL_692634\|2020-07 | .................... | .................... | ..................... | .................... | .................... | .................... |
| V | 1740 | hCoV-19/Japan/PG-4410/2020\|EPI_ISL_692635\|2020-07 | .................... | .................... | ..................... | .................... | .................... | .................... |
| V | 1741 | hCoV-19/Japan/PG-4411/2020\|EPI_ISL_692636\|2020-07 | .................... | .................... | ..................... | .................... | .................... | .................... |
| V | 1742 | hCoV-19/Japan/PG-4412/2020\|EPI_ISL_692637\|2020-07 | .................... | .................... | ..................... | .................... | .................... | .................... |
| V | 1743 | hCoV-19/Japan/PG-4413/2020\|EPI_ISL_692638\|2020-07 | .................... | .................... | ..................... | .................... | .................... | .................... |
| V | 1744 | hCoV-19/Japan/PG-4414/2020\|EPI_ISL_692639\|2020-07 | .................... | .................... | ..................... | .................... | .................... | .................... |
| V | 1745 | hCoV-19/Japan/PG-4415/2020\|EPI_ISL_692640\|2020-07 | .................... | .................... | ..................... | .................... | .................... | .................... |
| V | 1746 | hCoV-19/Japan/PG-4416/2020\|EPI_ISL_692641\|2020-07 | .................... | .................... | ..................... | .................... | .................... | .................... |
| V | 1747 | hCoV-19/Japan/PG-4417/2020\|EPI_ISL_692642\|2020-07 | .................... | .................... | ..................... | .................... | .................... | .................... |
| V | 1748 | hCoV-19/Japan/PG-4418/2020\|EPI_ISL_692643\|2020-07 | .................... | .................... | ..................... | .................... | .................... | .................... |
| V | 1749 | hCoV-19/Japan/PG-4419/2020\|EPI_ISL_692644\|2020-08 | .................... | .................... | ..................... | .................... | .................... | .................... |
| V | 1750 | hCoV-19/Japan/PG-4420/2020\|EPI_ISL_692645\|2020-08 | .................... | .................... | ..................... | .................... | .................... | .................... |
| V | 1751 | hCoV-19/Japan/PG-4421/2020\|EPI_ISL_692646\|2020-08 | .................... | .................... | ..................... | .................... | .................... | .................... |
| V | 1752 | hCoV-19/Japan/PG-4422/2020\|EPI_ISL_692647\|2020-08 | .................... | .................... | ..................... | .................... | .................... | .................... |
| V | 1753 | hCoV-19/Japan/PG-4423/2020\|EPI_ISL_692648\|2020-08 | .................... | .................... | ..................... | .................... | .................... | .................... |
| V | 1754 | hCoV-19/Japan/PG-4424/2020\|EPI_ISL_692649\|2020-08 | .................... | .................... | ..................... | .................... | .................... | .................... |
| V | 1755 | hCoV-19/Switzerland/AG-ETHZ-370282/2020\|EPI_ISL_69 | .................... | .................... | ..................... | .................... | .................... | .................... |
| V | 1756 | hCoV-19/Switzerland/BS-ETHZ-370312/2020\|EPI_ISL_69 | .................... | .................... | ..................... | .................... | .................... | .................... |
| V | 1757 | hCoV-19/Switzerland/BS-ETHZ-370330/2020\|EPI_ISL_69 | .................... | .................... | ..................... | .................... | .................... | .................... |
| V | 1758 | hCoV-19/Switzerland/BS-ETHZ-370334/2020\|EPI_ISL_69 | .................... | .................... | ..................... | .................... | .................... | .................... |
| V | 1759 | hCoV-19/Switzerland/BS-ETHZ-370356/2020\|EPI_ISL_69 | .................... | .................... | ..................... | .................... | .................... | .................... |
| V | 1760 | hCoV-19/Switzerland/BS-ETHZ-370358/2020\|EPI_ISL_69 | .................... | .................... | ..................... | .................... | .................... | .................... |
| V | 1761 | hCoV-19/Switzerland/BS-ETHZ-370372/2020\|EPI_ISL_69 | .................... | .................... | ..................... | .................... | .................... | .................... |
| V | 1762 | hCoV-19/Switzerland/ZH-ETHZ-370446/2020\|EPI_ISL_69 | .................... | .................... | .......K............. | .................... | .................... | .................... |
| V | 1763 | hCoV-19/Switzerland/GE-ETHZ-370447/2020\|EPI_ISL_69 | .................... | .................... | ..................... | .................... | .................... | .................... |
| V | 1764 | hCoV-19/Switzerland/VD-ETHZ-370448/2020\|EPI_ISL_69 | .................... | .................... | ..................... | .................... | .................... | .................... |
| V | 1765 | hCoV-19/Switzerland/VD-ETHZ-370449/2020\|EPI_ISL_69 | .................... | .................... | ..................... | .................... | ..............Y..... | .................... |
| V | 1766 | hCoV-19/Switzerland/SH-ETHZ-370450/2020\|EPI_ISL_69 | .................... | .A.................. | ..................... | .................... | .................... | .................... |
| V | 1767 | hCoV-19/Switzerland/SH-ETHZ-370452/2020\|EPI_ISL_69 | .................... | .................... | ..................... | .................... | .................... | .................... |
| V | 1768 | hCoV-19/Switzerland/SH-ETHZ-370453/2020\|EPI_ISL_69 | .................... | .................... | ..................... | .................... | .................... | .................... |
| V | 1769 | hCoV-19/Switzerland/SH-ETHZ-370454/2020\|EPI_ISL_69 | .................... | .A.................. | ..................... | .................... | .................... | .................... |
| V | 1770 | hCoV-19/Switzerland/VD-ETHZ-370455/2020\|EPI_ISL_69 | .................... | .................... | ..................... | .................... | ......K............. | .................... |
| V | 1771 | hCoV-19/Switzerland/SH-ETHZ-370456/2020\|EPI_ISL_69 | .................... | .................... | ..................... | .................... | .................... | .................... |
| V | 1772 | hCoV-19/Switzerland/SH-ETHZ-370458/2020\|EPI_ISL_69 | .................... | .A.................. | ..................... | .................... | .................... | .................... |
| V | 1773 | hCoV-19/Switzerland/SH-ETHZ-370460/2020\|EPI_ISL_69 | .................... | .................... | ..................... | .................... | .......S............ | .................... |
| V | 1774 | hCoV-19/Switzerland/SH-ETHZ-370461/2020\|EPI_ISL_69 | .................... | .................... | ..................... | .................... | .................... | .................... |
| V | 1775 | hCoV-19/Switzerland/SH-ETHZ-370462/2020\|EPI_ISL_69 | .................... | .................... | ..................... | .................... | .................... | .................... |
| V | 1776 | hCoV-19/Switzerland/SH-ETHZ-370463/2020\|EPI_ISL_69 | .................... | .................... | ..................... | .................... | .................... | .................... |
| V | 1777 | hCoV-19/Switzerland/SH-ETHZ-370464/2020\|EPI_ISL_69 | .................... | .................... | ..................... | .................... | .................... | .................... |
| V | 1778 | hCoV-19/Switzerland/BL-ETHZ-370465/2020\|EPI_ISL_69 | .................... | .................... | ..................... | .................... | ......K............. | .................... |
| V | 1779 | hCoV-19/Switzerland/TG-ETHZ-370466/2020\|EPI_ISL_69 | .................... | .................... | ..................... | .................... | .................... | .................... |
| V | 1780 | hCoV-19/Switzerland/ZH-ETHZ-370480/2020\|EPI_ISL_69 | .................... | .................... | ..................... | .................... | .................... | .................... |
| V | 1781 | hCoV-19/Switzerland/BE-ETHZ-370481/2020\|EPI_ISL_69 | .................... | .................... | ..................... | .................... | .................... | .................... |
| V | 1782 | hCoV-19/Switzerland/ZH-ETHZ-370482/2020\|EPI_ISL_69 | .................... | .................... | ..................... | .................... | .................... | .................... |
| V | 1783 | hCoV-19/Switzerland/BE-ETHZ-370484/2020\|EPI_ISL_69 | .................... | .................... | ..................... | .................... | .................... | .................... |
| V | 1784 | hCoV-19/Switzerland/BE-ETHZ-370485/2020\|EPI_ISL_69 | .................... | .................... | ..................... | .................... | .................... | .................... |
| V | 1785 | hCoV-19/Switzerland/BE-ETHZ-370487/2020\|EPI_ISL_69 | .................... | .................... | ..................... | .................... | .................... | .................... |
| V | 1786 | hCoV-19/Switzerland/BE-ETHZ-370488/2020\|EPI_ISL_69 | .................... | .................... | ..................... | .................... | .................... | .................... |
| V | 1787 | hCoV-19/Switzerland/VS-ETHZ-370489/2020\|EPI_ISL_69 | .................... | .................... | ..................... | .................... | .................... | .................... |
| V | 1788 | hCoV-19/Switzerland/VD-ETHZ-370490/2020\|EPI_ISL_69 | .................... | .................... | ..................... | .................... | .................... | .................... |
| V | 1789 | hCoV-19/Switzerland/VD-ETHZ-370491/2020\|EPI_ISL_69 | .................... | .................... | ..................... | .................... | .................... | .................... |
| V | 1790 | hCoV-19/Switzerland/VD-ETHZ-370492/2020\|EPI_ISL_69 | .................... | .................... | ..................... | .................... | .................... | .................... |
| V | 1791 | hCoV-19/Switzerland/GE-ETHZ-370493/2020\|EPI_ISL_69 | .................... | .................... | ..................... | .................... | .................... | .................... |
| V | 1792 | hCoV-19/Switzerland/GE-ETHZ-370494/2020\|EPI_ISL_69 | .................... | .................... | ..................... | .................... | .................... | .................... |
| V | 1793 | hCoV-19/Japan/PG-0305/2020\|EPI_ISL_684158\|2020-03 | .................... | .................... | ..................... | .................... | .................... | .................... |
| V | 1794 | hCoV-19/Japan/PG-0330/2020\|EPI_ISL_684159\|2020-03 | .................... | .................... | ..................... | .................... | .................... | .................... |
| V | 1795 | hCoV-19/Japan/PG-0335/2020\|EPI_ISL_684160\|2020-03 | .................... | .................... | ..................... | .................... | .................... | .................... |
| V | 1796 | hCoV-19/Japan/PG-0341/2020\|EPI_ISL_684161\|2020-03 | .................... | .................... | ..................... | .................... | .................... | .................... |
| V | 1797 | hCoV-19/Japan/PG-0358/2020\|EPI_ISL_684162\|2020-03 | .................... | .................... | ..................... | .................... | .................... | .................... |
| V | 1798 | hCoV-19/Japan/PG-0360/2020\|EPI_ISL_684163\|2020-02 | .................... | .................... | ..................... | .................... | .................... | .................... |
| V | 1799 | hCoV-19/Japan/PG-0380/2020\|EPI_ISL_684164\|2020-02 | .................... | .................... | ..................... | .................... | .................... | .................... |
| V | 1800 | hCoV-19/Japan/PG-0428/2020\|EPI_ISL_684166\|2020-04 | .................... | .................... | ..................... | .................... | .................... | .................... |
| V | 1801 | hCoV-19/Japan/PG-0431/2020\|EPI_ISL_684167\|2020-04 | .................... | .................... | ..................... | .................... | .................... | .................... |
| V | 1802 | hCoV-19/Japan/PG-0438/2020\|EPI_ISL_684168\|2020-03 | .................... | .................... | ..................... | .................... | .................... | .................... |
| V | 1803 | hCoV-19/Japan/PG-0447/2020\|EPI_ISL_684170\|2020-03 | .................... | .................... | ..................... | .................... | .................... | .................... |
| V | 1804 | hCoV-19/Japan/PG-0451/2020\|EPI_ISL_684171\|2020-03 | .................... | .................... | ..................... | .................... | .................... | .................... |
| V | 1805 | hCoV-19/Japan/PG-0456/2020\|EPI_ISL_684172\|2020-03 | .................... | .................... | ..................... | .................... | .................... | .................... |
| V | 1806 | hCoV-19/Japan/PG-0467/2020\|EPI_ISL_684173\|2020-03 | .................... | .................... | ..................... | .................... | .................... | .................... |
| V | 1807 | hCoV-19/Japan/PG-0479/2020\|EPI_ISL_684174\|2020-04 | .................... | .................... | ..................... | .................... | .................... | .................... |
| V | 1808 | hCoV-19/Japan/PG-0510/2020\|EPI_ISL_684175\|2020-04 | .................... | .................... | ..................... | .................... | .................... | .................... |
| V | 1809 | hCoV-19/Japan/PG-0535/2020\|EPI_ISL_684176\|2020-04 | .................... | .................... | ..................... | .................... | .................... | .................... |
| V | 1810 | hCoV-19/Japan/PG-0540/2020\|EPI_ISL_684177\|2020-04 | .................... | .................... | ..................... | .................... | .................... | .................... |
| V | 1811 | hCoV-19/Japan/PG-0548/2020\|EPI_ISL_684178\|2020-04 | .................... | .................... | ..................... | .................... | .................... | .................... |
| V | 1812 | hCoV-19/Japan/PG-0549/2020\|EPI_ISL_684179\|2020-04 | .................... | .................... | ..................... | .................... | .................... | .................... |
| V | 1813 | hCoV-19/Japan/PG-0551/2020\|EPI_ISL_684180\|2020-04 | .................... | .................... | ..................... | .................... | .................... | .................... |
| V | 1814 | hCoV-19/Japan/PG-0553/2020\|EPI_ISL_684181\|2020-04 | .................... | .................... | ..................... | .................... | .................... | .................... |

| | | | | | | | | |
|---|---|---|---|---|---|---|---|---|
| V | 1815 | hCoV-19/Japan/PG-0554/2020\|EPI_ISL_684182\|2020-04 | .................... | .................... | ..................... | .................... | .................... | .................... |
| V | 1816 | hCoV-19/Japan/PG-0558/2020\|EPI_ISL_684183\|2020-04 | .................... | .................... | ..................... | .................... | .................... | .................... |
| V | 1817 | hCoV-19/Japan/PG-0562/2020\|EPI_ISL_684184\|2020-04 | .................... | .................... | ..................... | .................... | .................... | .................... |
| V | 1818 | hCoV-19/Japan/PG-0565/2020\|EPI_ISL_684185\|2020-04 | .................... | .................... | ..................... | .................... | .................... | .................... |
| V | 1819 | hCoV-19/Japan/PG-0567/2020\|EPI_ISL_684186\|2020-04 | .................... | .................... | ..................... | .................... | .................... | .................... |
| V | 1820 | hCoV-19/Japan/PG-0568/2020\|EPI_ISL_684187\|2020-04 | .................... | .................... | ..................... | .................... | .................... | .................... |
| V | 1821 | hCoV-19/Japan/PG-0569/2020\|EPI_ISL_684188\|2020-04 | .................... | .................... | ..................... | .................... | .................... | .................... |
| V | 1822 | hCoV-19/Japan/PG-0570/2020\|EPI_ISL_684189\|2020-04 | .................... | .................... | ..................... | .................... | .................... | .................... |
| V | 1823 | hCoV-19/Japan/PG-0571/2020\|EPI_ISL_684190\|2020-04 | .................... | .................... | ..................... | .................... | .................... | .................... |
| V | 1824 | hCoV-19/Japan/PG-0574/2020\|EPI_ISL_684191\|2020-04 | .................... | .................... | ..................... | .................... | .................... | .................... |
| V | 1825 | hCoV-19/Japan/PG-0575/2020\|EPI_ISL_684192\|2020-04 | .................... | .................... | ..................... | .................... | .................... | .................... |
| V | 1826 | hCoV-19/Japan/PG-0577/2020\|EPI_ISL_684193\|2020-04 | .................... | .................... | ..................... | .................... | .................... | .................... |
| V | 1827 | hCoV-19/Japan/PG-0578/2020\|EPI_ISL_684194\|2020-04 | .................... | .................... | ..................... | .................... | .................... | .................... |
| V | 1828 | hCoV-19/Japan/PG-0580/2020\|EPI_ISL_684195\|2020-04 | .................... | .................... | ..................... | .................... | .................... | .................... |
| V | 1829 | hCoV-19/Japan/PG-0581/2020\|EPI_ISL_684196\|2020-04 | .................... | .................... | ..................... | .................... | .................... | .................... |
| V | 1830 | hCoV-19/Japan/PG-0583/2020\|EPI_ISL_684197\|2020-04 | .................... | .................... | ..................... | .................... | .................... | .................... |
| V | 1831 | hCoV-19/Japan/PG-0584/2020\|EPI_ISL_684198\|2020-04 | .................... | .................... | ..................... | .................... | .................... | .................... |
| V | 1832 | hCoV-19/Japan/PG-0585/2020\|EPI_ISL_684199\|2020-04 | .................... | .................... | ..................... | .................... | .................... | .................... |
| V | 1833 | hCoV-19/Japan/PG-1883/2020\|EPI_ISL_685489\|2020-05 | .................... | .................... | ..................... | .................... | .................... | .................... |
| V | 1834 | hCoV-19/Japan/PG-1884/2020\|EPI_ISL_685490\|2020-05 | .................... | .................... | ..................... | .................... | .................... | .................... |
| V | 1835 | hCoV-19/Japan/PG-1885/2020\|EPI_ISL_685491\|2020-05 | .................... | .................... | ..................... | .................... | .................... | .................... |
| V | 1836 | hCoV-19/Japan/PG-1886/2020\|EPI_ISL_685492\|2020-05 | .................... | .................... | ..................... | .................... | .................... | .................... |
| V | 1837 | hCoV-19/Japan/PG-1887/2020\|EPI_ISL_685493\|2020-05 | .................... | .................... | ..................... | .................... | .................... | .................... |
| V | 1838 | hCoV-19/Japan/PG-1888/2020\|EPI_ISL_685494\|2020-05 | .................... | .................... | ..................... | .................... | .................... | .................... |
| V | 1839 | hCoV-19/Japan/PG-1889/2020\|EPI_ISL_685495\|2020-05 | .................... | .................... | ..................... | .................... | .................... | .................... |
| V | 1840 | hCoV-19/Japan/PG-1890/2020\|EPI_ISL_685496\|2020-05 | .................... | .................... | ..................... | .................... | .................... | .................... |
| V | 1841 | hCoV-19/Japan/PG-1891/2020\|EPI_ISL_685497\|2020-05 | .................... | .................... | ..................... | .................... | .................... | .................... |
| V | 1842 | hCoV-19/Japan/PG-1892/2020\|EPI_ISL_685498\|2020-04 | .................... | .................... | ..................... | .................... | .................... | .................... |
| V | 1843 | hCoV-19/Japan/PG-1893/2020\|EPI_ISL_685499\|2020-04 | .................... | .................... | ..................... | .................... | .................... | .................... |
| V | 1844 | hCoV-19/Japan/PG-5885/2020\|EPI_ISL_689000\|2020-08 | .................... | .................... | ..................... | .................... | .................... | .................... |
| V | 1845 | hCoV-19/Japan/PG-5886/2020\|EPI_ISL_689001\|2020-08 | .................... | .................... | ..................... | .................... | .................... | .................... |
| V | 1846 | hCoV-19/Japan/PG-5887/2020\|EPI_ISL_689002\|2020-08 | .................... | .................... | ..................... | .................... | .................... | .................... |
| V | 1847 | hCoV-19/Japan/PG-5888/2020\|EPI_ISL_689003\|2020-08 | .................... | .................... | ..................... | .................... | .................... | .................... |
| V | 1848 | hCoV-19/Japan/PG-5889/2020\|EPI_ISL_689004\|2020-08 | .................... | .................... | ..................... | .................... | .................... | .................... |
| V | 1849 | hCoV-19/Japan/PG-5890/2020\|EPI_ISL_689005\|2020-08 | .................... | .................... | ..................... | .................... | .................... | .................... |
| V | 1850 | hCoV-19/Japan/PG-5891/2020\|EPI_ISL_689006\|2020-08 | .................... | .................... | ..................... | .................... | .................... | .................... |
| V | 1851 | hCoV-19/Japan/PG-5892/2020\|EPI_ISL_689007\|2020-08 | .................... | .................... | ..................... | .................... | .................... | .................... |
| V | 1852 | hCoV-19/Japan/PG-5893/2020\|EPI_ISL_689008\|2020-08 | .................... | .................... | ..................... | .................... | .................... | .................... |
| V | 1853 | hCoV-19/Japan/PG-5894/2020\|EPI_ISL_689009\|2020-08 | .................... | .................... | ..................... | .................... | .................... | .................... |
| V | 1854 | hCoV-19/Japan/PG-5895/2020\|EPI_ISL_689010\|2020-08 | .................... | .................... | ..................... | .................... | .................... | .................... |
| V | 1855 | hCoV-19/Japan/PG-5896/2020\|EPI_ISL_689011\|2020-08 | .................... | .................... | ..................... | .................... | .................... | .................... |
| V | 1856 | hCoV-19/Japan/PG-5897/2020\|EPI_ISL_689012\|2020-08 | .................... | .................... | ..................... | .................... | .................... | .................... |
| V | 1857 | hCoV-19/Japan/PG-5898/2020\|EPI_ISL_689013\|2020-08 | .................... | .................... | ..................... | .................... | .................... | .................... |
| V | 1858 | hCoV-19/Japan/PG-5899/2020\|EPI_ISL_689014\|2020-08 | .................... | .................... | ..................... | .................... | .................... | .................... |
| V | 1859 | hCoV-19/Japan/PG-5900/2020\|EPI_ISL_689015\|2020-08 | .................... | .................... | ..................... | .................... | .................... | .................... |
| V | 1860 | hCoV-19/Japan/PG-5901/2020\|EPI_ISL_689016\|2020-08 | .................... | .................... | ..................... | .................... | .................... | .................... |
| V | 1861 | hCoV-19/Japan/PG-5902/2020\|EPI_ISL_689017\|2020-08 | .................... | .................... | ..................... | .................... | .................... | .................... |
| V | 1862 | hCoV-19/Japan/PG-5903/2020\|EPI_ISL_689018\|2020-08 | .................... | .................... | ..................... | .................... | .................... | .................... |
| V | 1863 | hCoV-19/Japan/PG-5904/2020\|EPI_ISL_689019\|2020-08 | .................... | .................... | ..................... | .................... | .................... | .................... |
| V | 1864 | hCoV-19/Japan/PG-5905/2020\|EPI_ISL_689020\|2020-08 | .................... | .................... | ..................... | .................... | .................... | .................... |
| V | 1865 | hCoV-19/Japan/PG-5906/2020\|EPI_ISL_689021\|2020-08 | .................... | .................... | ..................... | .................... | .................... | .................... |
| V | 1866 | hCoV-19/Japan/PG-5907/2020\|EPI_ISL_689022\|2020-08 | .................... | .................... | ..................... | .................... | .................... | .................... |
| V | 1867 | hCoV-19/Japan/PG-5908/2020\|EPI_ISL_689023\|2020-08 | .................... | .................... | ..................... | .................... | .................... | .................... |
| V | 1868 | hCoV-19/Japan/PG-5909/2020\|EPI_ISL_689024\|2020-08 | .................... | .................... | ..................... | .................... | .................... | .................... |
| V | 1869 | hCoV-19/Japan/PG-5910/2020\|EPI_ISL_689025\|2020-08 | .................... | .................... | ..................... | .................... | .................... | .................... |
| V | 1870 | hCoV-19/Japan/PG-5911/2020\|EPI_ISL_689026\|2020-08 | .................... | .................... | ..................... | .................... | .................... | .................... |
| V | 1871 | hCoV-19/Japan/PG-5912/2020\|EPI_ISL_689027\|2020-08 | .................... | .................... | ..................... | .................... | .................... | .................... |
| V | 1872 | hCoV-19/Japan/PG-5913/2020\|EPI_ISL_689028\|2020-08 | .................... | .................... | ..................... | .................... | .................... | .................... |
| V | 1873 | hCoV-19/Japan/PG-5914/2020\|EPI_ISL_689029\|2020-08 | .................... | .................... | ..................... | .................... | .................... | .................... |
| V | 1874 | hCoV-19/Japan/PG-5915/2020\|EPI_ISL_689030\|2020-08 | .................... | .................... | ..................... | .................... | .................... | .................... |
| V | 1875 | hCoV-19/Japan/PG-5916/2020\|EPI_ISL_689031\|2020-08 | .................... | .................... | ..................... | .................... | .................... | .................... |
| V | 1876 | hCoV-19/Japan/PG-5917/2020\|EPI_ISL_689032\|2020-08 | .................... | .................... | ..................... | .................... | .................... | .................... |
| V | 1877 | hCoV-19/Japan/PG-5918/2020\|EPI_ISL_689033\|2020-08 | .................... | .................... | ..................... | .................... | .................... | .................... |
| V | 1878 | hCoV-19/Japan/PG-5919/2020\|EPI_ISL_689034\|2020-08 | .................... | .................... | ..................... | .................... | .................... | .................... |
| V | 1879 | hCoV-19/Japan/PG-5920/2020\|EPI_ISL_689035\|2020-08 | .................... | .................... | ..................... | .................... | .................... | .................... |
| V | 1880 | hCoV-19/Japan/PG-5921/2020\|EPI_ISL_689036\|2020-08 | .................... | .................... | ..................... | .................... | .................... | .................... |
| V | 1881 | hCoV-19/Japan/PG-5922/2020\|EPI_ISL_689037\|2020-08 | .................... | .................... | ..................... | .................... | .................... | .................... |
| V | 1882 | hCoV-19/Japan/PG-5923/2020\|EPI_ISL_689038\|2020-08 | .................... | .................... | ..................... | .................... | .................... | .................... |
| V | 1883 | hCoV-19/Japan/PG-5924/2020\|EPI_ISL_689039\|2020-08 | .................... | .................... | ..................... | .................... | .................... | .................... |
| V | 1884 | hCoV-19/Japan/PG-5925/2020\|EPI_ISL_689040\|2020-08 | .................... | .................... | ..................... | .................... | .................... | .................... |
| V | 1885 | hCoV-19/Japan/PG-5926/2020\|EPI_ISL_689041\|2020-08 | .................... | .................... | ..................... | .................... | .................... | .................... |
| V | 1886 | hCoV-19/Japan/PG-5927/2020\|EPI_ISL_689042\|2020-08 | .................... | .................... | ..................... | .................... | .................... | .................... |
| V | 1887 | hCoV-19/Japan/PG-5928/2020\|EPI_ISL_689043\|2020-08 | .................... | .................... | ..................... | .................... | .................... | .................... |
| V | 1888 | hCoV-19/Japan/PG-5929/2020\|EPI_ISL_689044\|2020-08 | .................... | .................... | ..................... | .................... | .................... | .................... |
| V | 1889 | hCoV-19/Japan/PG-5930/2020\|EPI_ISL_689045\|2020-08 | .................... | .................... | ..................... | .................... | .................... | .................... |
| V | 1890 | hCoV-19/Japan/PG-5931/2020\|EPI_ISL_689046\|2020-08 | .................... | .................... | ..................... | .................... | .................... | .................... |
| V | 1891 | hCoV-19/Japan/PG-5932/2020\|EPI_ISL_689047\|2020 | .................... | .................... | ..................... | .................... | .................... | .................... |
| V | 1892 | hCoV-19/Japan/PG-5933/2020\|EPI_ISL_689048\|2020 | .................... | .................... | ..................... | .................... | .................... | .................... |
| V | 1893 | hCoV-19/Japan/PG-5934/2020\|EPI_ISL_689049\|2020 | .................... | .................... | ..................... | .................... | .................... | .................... |
| V | 1894 | hCoV-19/Japan/PG-4879/2020\|EPI_ISL_690000\|2020-08 | .................... | .................... | ..................... | .................... | .................... | .................... |

| | | | | | | | | |
|---|---|---|---|---|---|---|---|---|
| V | 1895 | hCoV-19/Japan/PG-4880/2020\|EPI_ISL_690001\|2020-08 | .................... | .................... | ..................... | .................... | .................... | .................... |
| V | 1896 | hCoV-19/Japan/PG-4881/2020\|EPI_ISL_690002\|2020-08 | .................... | .................... | ..................... | .................... | .................... | .................... |
| V | 1897 | hCoV-19/Japan/PG-4882/2020\|EPI_ISL_690003\|2020-08 | .................... | .................... | ..................... | .................... | .................... | .................... |
| V | 1898 | hCoV-19/Japan/PG-4883/2020\|EPI_ISL_690004\|2020-08 | .................... | .................... | ..................... | .................... | .................... | .................... |
| V | 1899 | hCoV-19/Japan/PG-4884/2020\|EPI_ISL_690005\|2020-08 | .................... | .................... | ..................... | .................... | .................... | .................... |
| V | 1900 | hCoV-19/Japan/PG-4885/2020\|EPI_ISL_690006\|2020-08 | .................... | .................... | ..................... | .................... | .................... | .................... |
| V | 1901 | hCoV-19/Japan/PG-4886/2020\|EPI_ISL_690007\|2020-08 | .................... | .................... | ..................... | .................... | .................... | .................... |
| V | 1902 | hCoV-19/Japan/PG-4887/2020\|EPI_ISL_690008\|2020-08 | .................... | .................... | ..................... | .................... | .................... | .................... |
| V | 1903 | hCoV-19/Japan/PG-4888/2020\|EPI_ISL_690009\|2020-08 | .................... | .................... | ..................... | .................... | .................... | .................... |
| V | 1904 | hCoV-19/Japan/PG-4889/2020\|EPI_ISL_690010\|2020-08 | .................... | .................... | ..................... | .................... | .................... | .................... |
| V | 1905 | hCoV-19/Japan/PG-4890/2020\|EPI_ISL_690011\|2020-08 | .................... | .................... | ..................... | .................... | .................... | .................... |
| V | 1906 | hCoV-19/Japan/PG-4891/2020\|EPI_ISL_690012\|2020-08 | .................... | .................... | ..................... | .................... | .................... | .................... |
| V | 1907 | hCoV-19/Japan/PG-4892/2020\|EPI_ISL_690013\|2020-08 | .................... | .................... | ..................... | .................... | .................... | .................... |
| V | 1908 | hCoV-19/Japan/PG-4893/2020\|EPI_ISL_690014\|2020-08 | .................... | .................... | ..................... | .................... | .................... | .................... |
| V | 1909 | hCoV-19/Japan/PG-4894/2020\|EPI_ISL_690015\|2020-08 | .................... | .................... | ..................... | .................... | .................... | .................... |
| V | 1910 | hCoV-19/Japan/PG-4895/2020\|EPI_ISL_690016\|2020-08 | .................... | .................... | ..................... | .................... | .................... | .................... |
| V | 1911 | hCoV-19/Japan/PG-4896/2020\|EPI_ISL_690017\|2020-08 | .................... | .................... | ..................... | .................... | .................... | .................... |
| V | 1912 | hCoV-19/Japan/PG-4897/2020\|EPI_ISL_690018\|2020-08 | .................... | .................... | ..................... | .................... | .................... | .................... |
| V | 1913 | hCoV-19/Japan/PG-4898/2020\|EPI_ISL_690019\|2020-08 | .................... | .................... | ..................... | .................... | .................... | .................... |
| V | 1914 | hCoV-19/Japan/PG-4899/2020\|EPI_ISL_690020\|2020-08 | .................... | .................... | ..................... | .................... | .................... | .................... |
| V | 1915 | hCoV-19/Japan/PG-4900/2020\|EPI_ISL_690021\|2020-08 | .................... | .................... | ..................... | .................... | .................... | .................... |
| V | 1916 | hCoV-19/Japan/PG-4901/2020\|EPI_ISL_690022\|2020-08 | .................... | .................... | ..................... | .................... | .................... | .................... |
| V | 1917 | hCoV-19/Japan/PG-4902/2020\|EPI_ISL_690023\|2020-08 | .................... | .................... | ..................... | .................... | .................... | .................... |
| V | 1918 | hCoV-19/Japan/PG-4903/2020\|EPI_ISL_690024\|2020-08 | .................... | .................... | ..................... | .................... | .................... | .................... |
| V | 1919 | hCoV-19/Japan/PG-4904/2020\|EPI_ISL_690025\|2020-08 | .................... | .................... | ..................... | .................... | .................... | .................... |
| V | 1920 | hCoV-19/Japan/PG-4905/2020\|EPI_ISL_690026\|2020-08 | .................... | .................... | ..................... | .................... | .................... | .................... |
| V | 1921 | hCoV-19/Japan/PG-4906/2020\|EPI_ISL_690027\|2020-08 | .................... | .................... | ..................... | .................... | .................... | .................... |
| V | 1922 | hCoV-19/Japan/PG-4907/2020\|EPI_ISL_690028\|2020-08 | .................... | .................... | ..................... | .................... | .................... | .................... |
| V | 1923 | hCoV-19/Japan/PG-4908/2020\|EPI_ISL_690029\|2020-07 | .................... | .................... | ..................... | .................... | .................... | .................... |
| V | 1924 | hCoV-19/Japan/PG-4909/2020\|EPI_ISL_690030\|2020-07 | .................... | .................... | ..................... | .................... | .................... | .................... |
| V | 1925 | hCoV-19/Japan/PG-4911/2020\|EPI_ISL_690031\|2020-07 | .................... | .................... | ..................... | .................... | .................... | .................... |
| V | 1926 | hCoV-19/Japan/PG-3200/2020\|EPI_ISL_691320\|2020-04 | .................... | .................... | ..................... | .................... | .................... | .................... |
| V | 1927 | hCoV-19/Japan/PG-3201/2020\|EPI_ISL_691321\|2020-04 | .................... | .................... | ..................... | .................... | .................... | .................... |
| V | 1928 | hCoV-19/Japan/PG-3202/2020\|EPI_ISL_691322\|2020-04 | .................... | .................... | ..................... | .................... | .................... | .................... |
| V | 1929 | hCoV-19/Japan/PG-3203/2020\|EPI_ISL_691323\|2020-04 | .................... | .................... | ..................... | .................... | .................... | .................... |
| V | 1930 | hCoV-19/Japan/PG-3204/2020\|EPI_ISL_691324\|2020-04 | .................... | .................... | ..................... | .................... | .................... | .................... |
| V | 1931 | hCoV-19/Japan/PG-3206/2020\|EPI_ISL_691325\|2020-04 | .................... | .................... | ..................... | .................... | .................... | .................... |
| V | 1932 | hCoV-19/Japan/PG-3208/2020\|EPI_ISL_691326\|2020-04 | .................... | .................... | ..................... | .................... | .................... | .................... |
| V | 1933 | hCoV-19/Japan/PG-3209/2020\|EPI_ISL_691327\|2020-04 | .................... | .................... | ..................... | .................... | .................... | .................... |
| V | 1934 | hCoV-19/Japan/PG-3210/2020\|EPI_ISL_691328\|2020-04 | .................... | .................... | ..................... | .................... | .................... | .................... |
| V | 1935 | hCoV-19/Japan/PG-3211/2020\|EPI_ISL_691329\|2020-04 | .................... | .................... | ..................... | .................... | .................... | .................... |
| V | 1936 | hCoV-19/Japan/PG-3212/2020\|EPI_ISL_691330\|2020-04 | .................... | .................... | ..................... | .................... | .................... | .................... |
| V | 1937 | hCoV-19/Japan/PG-3213/2020\|EPI_ISL_691331\|2020-04 | .................... | .................... | ..................... | .................... | .................... | .................... |
| V | 1938 | hCoV-19/Japan/PG-3214/2020\|EPI_ISL_691332\|2020-04 | .................... | .................... | ..................... | .................... | .................... | .................... |
| V | 1939 | hCoV-19/Japan/PG-3215/2020\|EPI_ISL_691333\|2020-04 | .................... | .................... | ..................... | .................... | .................... | .................... |
| V | 1940 | hCoV-19/Japan/PG-3216/2020\|EPI_ISL_691334\|2020-04 | .................... | .................... | ..................... | .................... | .................... | .................... |
| V | 1941 | hCoV-19/Japan/PG-3217/2020\|EPI_ISL_691335\|2020-04 | .................... | .................... | ..................... | .................... | .................... | .................... |
| V | 1942 | hCoV-19/Japan/PG-3218/2020\|EPI_ISL_691336\|2020-04 | .................... | .................... | ..................... | .................... | .................... | .................... |
| V | 1943 | hCoV-19/Japan/PG-3219/2020\|EPI_ISL_691337\|2020-04 | .................... | .................... | ..................... | .................... | .................... | .................... |
| V | 1944 | hCoV-19/Japan/PG-3220/2020\|EPI_ISL_691338\|2020-04 | .................... | .................... | ..................... | .................... | .................... | .................... |
| V | 1945 | hCoV-19/Japan/PG-3221/2020\|EPI_ISL_691339\|2020-04 | .................... | .................... | ..................... | .................... | .................... | .................... |
| V | 1946 | hCoV-19/Japan/PG-3222/2020\|EPI_ISL_691340\|2020-04 | .................... | .................... | ..................... | .................... | .................... | .................... |
| V | 1947 | hCoV-19/Japan/PG-3224/2020\|EPI_ISL_691341\|2020-04 | .................... | .................... | ..................... | .................... | .................... | .................... |
| V | 1948 | hCoV-19/Japan/PG-3225/2020\|EPI_ISL_691342\|2020-04 | .................... | .................... | ..................... | .................... | .................... | .................... |
| V | 1949 | hCoV-19/Japan/PG-3226/2020\|EPI_ISL_691343\|2020-04 | .................... | .................... | ..................... | .................... | .................... | .................... |
| V | 1950 | hCoV-19/Japan/PG-3227/2020\|EPI_ISL_691344\|2020-04 | .................... | .................... | ..................... | .................... | .................... | .................... |
| V | 1951 | hCoV-19/Japan/PG-3228/2020\|EPI_ISL_691345\|2020-04 | .................... | .................... | ..................... | .................... | .................... | .................... |
| V | 1952 | hCoV-19/Japan/PG-3229/2020\|EPI_ISL_691346\|2020-04 | .................... | .................... | ..................... | .................... | .................... | .................... |
| V | 1953 | hCoV-19/Japan/PG-3230/2020\|EPI_ISL_691347\|2020-04 | .................... | .................... | ..................... | .................... | .................... | .................... |
| V | 1954 | hCoV-19/Japan/PG-3231/2020\|EPI_ISL_691348\|2020-04 | .................... | .................... | ..................... | .................... | .................... | .................... |
| V | 1955 | hCoV-19/Japan/PG-3232/2020\|EPI_ISL_691349\|2020-04 | .................... | .................... | ..................... | .................... | .................... | .................... |
| V | 1956 | hCoV-19/Japan/PG-3234/2020\|EPI_ISL_691350\|2020-04 | .................... | .................... | ..................... | .................... | .................... | .................... |
| V | 1957 | hCoV-19/Japan/PG-3235/2020\|EPI_ISL_691351\|2020-04 | .................... | .................... | ..................... | .................... | .................... | .................... |
| V | 1958 | hCoV-19/Japan/PG-3236/2020\|EPI_ISL_691352\|2020-04 | .................... | .................... | ..................... | .................... | .................... | .................... |
| V | 1959 | hCoV-19/Japan/PG-3237/2020\|EPI_ISL_691353\|2020-04 | .................... | .................... | ..................... | .................... | .................... | .................... |
| V | 1960 | hCoV-19/Japan/PG-3238/2020\|EPI_ISL_691354\|2020-04 | .................... | .................... | ..................... | .................... | .................... | .................... |
| V | 1961 | hCoV-19/Japan/PG-3239/2020\|EPI_ISL_691355\|2020-04 | .................... | .................... | ..................... | .................... | .................... | .................... |
| V | 1962 | hCoV-19/Japan/PG-3240/2020\|EPI_ISL_691356\|2020-04 | .................... | .................... | ..................... | .................... | .................... | .................... |
| V | 1963 | hCoV-19/Japan/PG-3241/2020\|EPI_ISL_691357\|2020-04 | .................... | .................... | ..................... | .................... | .................... | .................... |
| V | 1964 | hCoV-19/Japan/PG-3242/2020\|EPI_ISL_691358\|2020-04 | .................... | .................... | ..................... | .................... | .................... | .................... |
| V | 1965 | hCoV-19/Japan/PG-3243/2020\|EPI_ISL_691359\|2020-04 | .................... | .................... | ..................... | .................... | .................... | .................... |
| V | 1966 | hCoV-19/Japan/PG-3244/2020\|EPI_ISL_691360\|2020-04 | .................... | .................... | ..................... | .................... | .................... | .................... |
| V | 1967 | hCoV-19/Japan/PG-3246/2020\|EPI_ISL_691361\|2020-04 | .................... | .................... | ..................... | .................... | .................... | .................... |
| V | 1968 | hCoV-19/Japan/PG-3247/2020\|EPI_ISL_691362\|2020-04 | .................... | .................... | ..................... | .................... | .................... | .................... |
| V | 1969 | hCoV-19/Japan/PG-4426/2020\|EPI_ISL_692650\|2020-07 | .................... | .................... | ..................... | .................... | .................... | .................... |
| V | 1970 | hCoV-19/Japan/PG-4427/2020\|EPI_ISL_692651\|2020-07 | .................... | .................... | ..................... | .................... | .................... | .................... |
| V | 1971 | hCoV-19/Japan/PG-4428/2020\|EPI_ISL_692652\|2020-07 | .................... | .................... | ..................... | .................... | .................... | .................... |
| V | 1972 | hCoV-19/Japan/PG-4429/2020\|EPI_ISL_692653\|2020-07 | .................... | .................... | ..................... | .................... | .................... | .................... |
| V | 1973 | hCoV-19/Japan/PG-4430/2020\|EPI_ISL_692654\|2020-07 | .................... | .................... | ..................... | .................... | .................... | .................... |
| V | 1974 | hCoV-19/Japan/PG-4431/2020\|EPI_ISL_692655\|2020-07 | .................... | .................... | ..................... | .................... | .................... | .................... |

| | | | | | | | | |
|---|---|---|---|---|---|---|---|---|
| V | 1975 | hCoV-19/Japan/PG-4433/2020\|EPI_ISL_692656\|2020-07 | ···················· | ···················· | ···················· | ···················· | ···················· | ···················· |
| V | 1976 | hCoV-19/Japan/PG-4434/2020\|EPI_ISL_692657\|2020-07 | ···················· | ···················· | ···················· | ···················· | ···················· | ···················· |
| V | 1977 | hCoV-19/Japan/PG-4435/2020\|EPI_ISL_692658\|2020-07 | ···················· | ···················· | ···················· | ···················· | ···················· | ···················· |
| V | 1978 | hCoV-19/Japan/PG-4436/2020\|EPI_ISL_692659\|2020-07 | ···················· | ···················· | ···················· | ···················· | ···················· | ···················· |
| V | 1979 | hCoV-19/Japan/PG-4437/2020\|EPI_ISL_692660\|2020-07 | ···················· | ···················· | ···················· | ···················· | ···················· | ···················· |
| V | 1980 | hCoV-19/Japan/PG-4438/2020\|EPI_ISL_692661\|2020-07 | ···················· | ···················· | ···················· | ···················· | ···················· | ···················· |
| V | 1981 | hCoV-19/Japan/PG-4439/2020\|EPI_ISL_692662\|2020-07 | ···················· | ···················· | ···················· | ···················· | ···················· | ···················· |
| V | 1982 | hCoV-19/Japan/PG-4440/2020\|EPI_ISL_692663\|2020-07 | ···················· | ···················· | ···················· | ···················· | ···················· | ···················· |
| V | 1983 | hCoV-19/Japan/PG-4441/2020\|EPI_ISL_692664\|2020-07 | ···················· | ···················· | ···················· | ···················· | ···················· | ···················· |
| V | 1984 | hCoV-19/Japan/PG-4442/2020\|EPI_ISL_692665\|2020-07 | ···················· | ···················· | ···················· | ···················· | ···················· | ···················· |
| V | 1985 | hCoV-19/Japan/PG-4443/2020\|EPI_ISL_692666\|2020-07 | ···················· | ···················· | ···················· | ···················· | ···················· | ···················· |
| V | 1986 | hCoV-19/Japan/PG-4444/2020\|EPI_ISL_692667\|2020-07 | ···················· | ···················· | ···················· | ···················· | ···················· | ···················· |
| V | 1987 | hCoV-19/Japan/PG-4445/2020\|EPI_ISL_692668\|2020-07 | ···················· | ···················· | ···················· | ···················· | ···················· | ···················· |
| V | 1988 | hCoV-19/Japan/PG-4446/2020\|EPI_ISL_692669\|2020-07 | ···················· | ···················· | ···················· | ···················· | ···················· | ···················· |
| V | 1989 | hCoV-19/Japan/PG-4447/2020\|EPI_ISL_692670\|2020-07 | ···················· | ···················· | ···················· | ···················· | ···················· | ···················· |
| V | 1990 | hCoV-19/Japan/PG-4448/2020\|EPI_ISL_692671\|2020-07 | ···················· | ···················· | ···················· | ···················· | ···················· | ···················· |
| V | 1991 | hCoV-19/Japan/PG-4449/2020\|EPI_ISL_692672\|2020-07 | ···················· | ···················· | ···················· | ···················· | ···················· | ···················· |
| V | 1992 | hCoV-19/Japan/PG-4450/2020\|EPI_ISL_692673\|2020-07 | ···················· | ···················· | ···················· | ···················· | ···················· | ···················· |
| V | 1993 | hCoV-19/Japan/PG-4451/2020\|EPI_ISL_692674\|2020-07 | ···················· | ···················· | ···················· | ···················· | ···················· | ···················· |
| V | 1994 | hCoV-19/Japan/PG-4452/2020\|EPI_ISL_692675\|2020-07 | ···················· | ···················· | ···················· | ···················· | ···················· | ···················· |
| V | 1995 | hCoV-19/Japan/PG-4453/2020\|EPI_ISL_692676\|2020-07 | ···················· | ···················· | ···················· | ···················· | ···················· | ···················· |
| V | 1996 | hCoV-19/Japan/PG-4454/2020\|EPI_ISL_692677\|2020-07 | ···················· | ···················· | ···················· | ···················· | ···················· | ···················· |
| V | 1997 | hCoV-19/Japan/PG-4455/2020\|EPI_ISL_692678\|2020-08 | ···················· | ···················· | ···················· | ···················· | ···················· | ···················· |
| V | 1998 | hCoV-19/Japan/PG-4456/2020\|EPI_ISL_692679\|2020-08 | ···················· | ···················· | ···················· | ···················· | ···················· | ···················· |
| V | 1999 | hCoV-19/Japan/PG-4457/2020\|EPI_ISL_692680\|2020-07 | ···················· | ···················· | ···················· | ···················· | ···················· | ···················· |
| V | 2000 | hCoV-19/Japan/PG-4458/2020\|EPI_ISL_692681\|2020-07 | ···················· | ···················· | ···················· | ···················· | ···················· | ···················· |
| V | 2002 | hCoV-19/Japan/PG-4460/2020\|EPI_ISL_692683\|2020-07 | ···················· | ···················· | ···················· | ···················· | ···················· | ···················· |
| V | 2003 | hCoV-19/Japan/PG-4461/2020\|EPI_ISL_692684\|2020-07 | ···················· | ···················· | ···················· | ···················· | ···················· | ···················· |
| V | 2004 | hCoV-19/Japan/PG-4462/2020\|EPI_ISL_692685\|2020-07 | ···················· | ···················· | ···················· | ···················· | ···················· | ···················· |
| V | 2005 | hCoV-19/Japan/PG-4463/2020\|EPI_ISL_692686\|2020-07 | ···················· | ···················· | ···················· | ···················· | ···················· | ···················· |
| V | 2006 | hCoV-19/Japan/PG-4464/2020\|EPI_ISL_692687\|2020-07 | ···················· | ···················· | ···················· | ···················· | ···················· | ···················· |
| V | 2007 | hCoV-19/Japan/PG-4465/2020\|EPI_ISL_692688\|2020-07 | ···················· | ···················· | ···················· | ···················· | ···················· | ···················· |
| V | 2008 | hCoV-19/Japan/PG-4466/2020\|EPI_ISL_692689\|2020-07 | ···················· | ···················· | ···················· | ···················· | ···················· | ···················· |
| V | 2009 | hCoV-19/Japan/PG-4467/2020\|EPI_ISL_692690\|2020-07 | ···················· | ···················· | ···················· | ···················· | ···················· | ···················· |
| V | 2010 | hCoV-19/Japan/PG-4468/2020\|EPI_ISL_692691\|2020-07 | ···················· | ···················· | ···················· | ···················· | ···················· | ···················· |
| V | 2011 | hCoV-19/Japan/PG-4469/2020\|EPI_ISL_692692\|2020-07 | ···················· | ···················· | ···················· | ···················· | ···················· | ···················· |
| V | 2012 | hCoV-19/Japan/PG-4470/2020\|EPI_ISL_692693\|2020-07 | ···················· | ···················· | ···················· | ···················· | ···················· | ···················· |
| V | 2013 | hCoV-19/Switzerland/BE-ETHZ-360090/2020\|EPI_ISL_69 | ···················· | ···················· | ···················· | ···················· | ···················· | ···················· |
| V | 2014 | hCoV-19/Switzerland/BE-ETHZ-360091/2020\|EPI_ISL_69 | ···················· | ···················· | ···················· | ···················· | ···················· | ···················· |
| V | 2015 | hCoV-19/Switzerland/BE-ETHZ-360092/2020\|EPI_ISL_69 | ···················· | ···················· | ···················· | ···················· | ···················· | ···················· |
| V | 2016 | hCoV-19/Switzerland/BE-ETHZ-360093/2020\|EPI_ISL_69 | ···················· | ···················· | ···················· | ···················· | ·······S············ | ···················· |
| V | 2017 | hCoV-19/Switzerland/BE-ETHZ-360094/2020\|EPI_ISL_69 | ···················· | ···················· | ···················· | ···················· | ···················· | ···················· |
| V | 2018 | hCoV-19/Switzerland/BE-ETHZ-360095/2020\|EPI_ISL_69 | ···················· | ···················· | ···················· | ···················· | ···················· | ···················· |
| V | 2019 | hCoV-19/Switzerland/BE-ETHZ-360097/2020\|EPI_ISL_69 | ···················· | ···················· | ···················· | ···················· | ···················· | ···················· |
| V | 2020 | hCoV-19/Switzerland/BE-ETHZ-360098/2020\|EPI_ISL_69 | ···················· | ···················· | ···················· | ···················· | ···················· | ···················· |
| V | 2021 | hCoV-19/Switzerland/GR-ETHZ-360100/2020\|EPI_ISL_69 | ···················· | ···················· | ···················· | ···················· | ···················· | ···················· |
| V | 2022 | hCoV-19/Switzerland/SZ-ETHZ-360101/2020\|EPI_ISL_69 | ···················· | ···················· | ···················· | ···················· | ···················· | ···················· |
| V | 2023 | hCoV-19/Switzerland/ZH-ETHZ-360117/2020\|EPI_ISL_69 | ···················· | ···················· | ···················· | ···················· | ···················· | ···················· |
| V | 2024 | hCoV-19/Switzerland/SZ-ETHZ-360118/2020\|EPI_ISL_69 | ···················· | ···················· | ···················· | ···················· | ···················· | ···················· |
| V | 2025 | hCoV-19/Switzerland/GE-ETHZ-360136/2020\|EPI_ISL_69 | ···················· | ···················· | ···················· | ···················· | ···················· | ···················· |
| V | 2026 | hCoV-19/Switzerland/BL-ETHZ-360138/2020\|EPI_ISL_69 | ···················· | ···················· | ···················· | ···················· | ···················· | ···················· |
| V | 2027 | hCoV-19/Switzerland/BE-ETHZ-360139/2020\|EPI_ISL_69 | ···················· | ···················· | ···················· | ···················· | ···········W········ | ···················· |
| V | 2028 | hCoV-19/Switzerland/BE-ETHZ-360140/2020\|EPI_ISL_69 | ···················· | ···················· | ···················· | ···················· | ···················· | ···················· |
| V | 2029 | hCoV-19/Switzerland/GE-ETHZ-370495/2020\|EPI_ISL_69 | ···················· | ···················· | ···················· | ···················· | ···················· | ···················· |
| V | 2030 | hCoV-19/Switzerland/GE-ETHZ-370496/2020\|EPI_ISL_69 | ···················· | ···················· | ···················· | ···················· | ···················· | ···················· |
| V | 2031 | hCoV-19/Switzerland/BE-ETHZ-370498/2020\|EPI_ISL_69 | ···················· | ·A·················· | ···················· | ···················· | ···················· | ···················· |
| V | 2032 | hCoV-19/Switzerland/BE-ETHZ-370499/2020\|EPI_ISL_69 | ···················· | ···················· | ···················· | ···················· | ···················· | ···················· |
| V | 2033 | hCoV-19/Switzerland/BE-ETHZ-370500/2020\|EPI_ISL_69 | ···················· | ···················· | ···················· | ···················· | ···················· | ···················· |
| V | 2034 | hCoV-19/Switzerland/BE-ETHZ-370501/2020\|EPI_ISL_69 | ···················· | ···················· | ···················· | ···················· | ···················· | ···················· |
| V | 2035 | hCoV-19/Switzerland/BE-ETHZ-370502/2020\|EPI_ISL_69 | ···················· | ···················· | ···················· | ···················· | ···················· | ···················· |
| V | 2036 | hCoV-19/Switzerland/BE-ETHZ-370503/2020\|EPI_ISL_69 | ···················· | ···················· | ···················· | ···················· | ···················· | ···················· |
| V | 2037 | hCoV-19/Switzerland/BE-ETHZ-370504/2020\|EPI_ISL_69 | ···················· | ···················· | ···················· | ···················· | ···················· | ···················· |
| V | 2038 | hCoV-19/Switzerland/BE-ETHZ-370505/2020\|EPI_ISL_69 | ···················· | ···················· | ···················· | ···················· | ···················· | ···················· |
| V | 2039 | hCoV-19/Switzerland/BE-ETHZ-370506/2020\|EPI_ISL_69 | ···················· | ···················· | ···················· | ···················· | ···················· | ···················· |
| V | 2040 | hCoV-19/Switzerland/VD-ETHZ-370507/2020\|EPI_ISL_69 | ···················· | ···················· | ···················· | ···················· | ···················· | ···················· |
| V | 2041 | hCoV-19/Switzerland/BE-ETHZ-370508/2020\|EPI_ISL_69 | ···················· | ···················· | ···················· | ···················· | ···················· | ···················· |
| V | 2042 | hCoV-19/Switzerland/BE-ETHZ-370509/2020\|EPI_ISL_69 | ···················· | ···················· | ···················· | ···················· | ···················· | ···················· |
| V | 2043 | hCoV-19/Switzerland/BE-ETHZ-370510/2020\|EPI_ISL_69 | ···················· | ···················· | ···················· | ···················· | ···················· | ···················· |
| V | 2044 | hCoV-19/Switzerland/ZH-ETHZ-370511/2020\|EPI_ISL_69 | ···················· | ···················· | ···················· | ···················· | ······K············· | ···················· |
| V | 2045 | hCoV-19/Switzerland/AG-ETHZ-370513/2020\|EPI_ISL_69 | ···················· | ···················· | ···················· | ···················· | ······K············· | ···················· |
| V | 2046 | hCoV-19/Switzerland/ZH-ETHZ-370515/2020\|EPI_ISL_69 | ···················· | ···················· | ···················· | ···················· | ···················· | ···················· |
| V | 2047 | hCoV-19/Switzerland/ZH-ETHZ-370516/2020\|EPI_ISL_69 | ···················· | ·A·················· | ···················· | ···················· | ···················· | ···················· |
| V | 2048 | hCoV-19/Switzerland/ZH-ETHZ-370517/2020\|EPI_ISL_69 | ···················· | ···············R···· | ···················· | ···················· | ···················· | ···················· |
| V | 2049 | hCoV-19/Japan/PG-0073/2020\|EPI_ISL_684110\|2020-02 | ···················· | ···················· | ···················· | ···················· | ···················· | ···················· |
| V | 2050 | hCoV-19/Japan/PG-0075/2020\|EPI_ISL_684111\|2020-02 | ···················· | ···················· | ···················· | ···················· | ···················· | ···················· |
| V | 2051 | hCoV-19/Japan/PG-0079/2020\|EPI_ISL_684113\|2020-02 | ···················· | ···················· | ···················· | ···················· | ···················· | ···················· |
| V | 2052 | hCoV-19/Japan/PG-0083/2020\|EPI_ISL_684114\|2020-03 | ···················· | ···················· | ···················· | ···················· | ···················· | ···················· |
| V | 2053 | hCoV-19/Japan/PG-0085/2020\|EPI_ISL_684115\|2020-03 | ···················· | ···················· | ···················· | ···················· | ···················· | ···················· |
| V | 2054 | hCoV-19/Japan/PG-0086/2020\|EPI_ISL_684116\|2020-03 | ···················· | ···················· | ···················· | ···················· | ···················· | ···················· |
| V | 2055 | hCoV-19/Japan/PG-0090/2020\|EPI_ISL_684118\|2020-03 | ···················· | ···················· | ···················· | ···················· | ···················· | ···················· |

| | | | | | | | | |
|---|---|---|---|---|---|---|---|---|
| V | 2056 | hCoV-19/Japan/PG-0093/2020\|EPI_ISL_684119\|2020-03 | .................... | .................... | ..................... | .................... | .................... | .................... |
| V | 2057 | hCoV-19/Japan/PG-0094/2020\|EPI_ISL_684120\|2020-03 | .................... | .................... | ..................... | .................... | .................... | .................... |
| V | 2058 | hCoV-19/Japan/PG-0096/2020\|EPI_ISL_684121\|2020-03 | .................... | .................... | ..................... | .................... | .................... | .................... |
| V | 2059 | hCoV-19/Japan/PG-0101/2020\|EPI_ISL_684122\|2020-03 | .................... | .................... | ..................... | .................... | .................... | .................... |
| V | 2060 | hCoV-19/Japan/PG-0102/2020\|EPI_ISL_684123\|2020-03 | .................... | .................... | ..................... | .................... | .................... | .................... |
| V | 2061 | hCoV-19/Japan/PG-0105/2020\|EPI_ISL_684124\|2020-03 | .................... | .................... | ..................... | .................... | .................... | .................... |
| V | 2062 | hCoV-19/Japan/PG-0107/2020\|EPI_ISL_684125\|2020-03 | .................... | .................... | ..................... | .................... | .................... | .................... |
| V | 2063 | hCoV-19/Japan/PG-0112/2020\|EPI_ISL_684126\|2020-03 | .................... | .................... | ..................... | .................... | .................... | .................... |
| V | 2064 | hCoV-19/Japan/PG-0126/2020\|EPI_ISL_684128\|2020-03 | .................... | .................... | ..................... | .................... | .................... | .................... |
| V | 2065 | hCoV-19/Japan/PG-0129/2020\|EPI_ISL_684129\|2020-03 | .................... | .................... | ..................... | .................... | .................... | .................... |
| V | 2066 | hCoV-19/Japan/PG-0142/2020\|EPI_ISL_684130\|2020-02 | .................... | .................... | ..................... | .................... | .................... | .................... |
| V | 2067 | hCoV-19/Japan/PG-0155/2020\|EPI_ISL_684132\|2020-03 | .................... | .................... | ..................... | .................... | .................... | .................... |
| V | 2068 | hCoV-19/Japan/PG-0156/2020\|EPI_ISL_684133\|2020-03 | .................... | .................... | ..................... | .................... | .................... | .................... |
| V | 2069 | hCoV-19/Japan/PG-0177/2020\|EPI_ISL_684136\|2020-03 | .................... | .................... | ..................... | .................... | .................... | .................... |
| V | 2070 | hCoV-19/Japan/PG-0199/2020\|EPI_ISL_684139\|2020-02 | .................... | .................... | ..................... | .................... | .................... | .................... |
| V | 2071 | hCoV-19/Japan/PG-0206/2020\|EPI_ISL_684142\|2020-03 | .................... | .................... | ..................... | .................... | .................... | .................... |
| V | 2072 | hCoV-19/Japan/PG-0220/2020\|EPI_ISL_684143\|2020-03 | .................... | .................... | ..................... | .................... | .................... | .................... |
| V | 2073 | hCoV-19/Japan/PG-0226/2020\|EPI_ISL_684145\|2020-03 | .................... | .................... | ..................... | .................... | .................... | .................... |
| V | 2074 | hCoV-19/Japan/PG-0228/2020\|EPI_ISL_684146\|2020-03 | .................... | .................... | ..................... | .................... | .................... | .................... |
| V | 2075 | hCoV-19/Japan/PG-0244/2020\|EPI_ISL_684149\|2020-03 | .................... | .................... | ..................... | .................... | .................... | .................... |
| V | 2076 | hCoV-19/Japan/PG-0257/2020\|EPI_ISL_684150\|2020-03 | .................... | .................... | ..................... | .................... | .................... | .................... |
| V | 2077 | hCoV-19/Japan/PG-0266/2020\|EPI_ISL_684151\|2020-03 | .................... | .................... | ..................... | .................... | .................... | .................... |
| V | 2078 | hCoV-19/Japan/PG-0270/2020\|EPI_ISL_684152\|2020-03 | .................... | .................... | ..................... | .................... | .................... | .................... |
| V | 2079 | hCoV-19/Japan/PG-0273/2020\|EPI_ISL_684153\|2020-03 | .................... | .................... | ..................... | .................... | .................... | .................... |
| V | 2080 | hCoV-19/Japan/PG-0278/2020\|EPI_ISL_684154\|2020-03 | .................... | .................... | ..................... | .................... | .................... | .................... |
| V | 2081 | hCoV-19/Japan/PG-0282/2020\|EPI_ISL_684155\|2020-03 | .................... | .................... | ..................... | .................... | .................... | .................... |
| V | 2082 | hCoV-19/Japan/PG-0288/2020\|EPI_ISL_684156\|2020-03 | .................... | .................... | ..................... | .................... | .................... | .................... |
| V | 2083 | hCoV-19/Japan/PG-0298/2020\|EPI_ISL_684157\|2020-03 | .................... | .................... | ..................... | .................... | .................... | .................... |
| V | 2084 | hCoV-19/Japan/PG-1794/2020\|EPI_ISL_685400\|2020-05 | .................... | .................... | ..................... | .................... | .................... | .................... |
| V | 2085 | hCoV-19/Japan/PG-1834/2020\|EPI_ISL_685440\|2020-04 | .................... | .................... | ..................... | .................... | .................... | .................... |
| V | 2086 | hCoV-19/Japan/PG-1835/2020\|EPI_ISL_685441\|2020-04 | .................... | .................... | ..................... | .................... | .................... | .................... |
| V | 2087 | hCoV-19/Japan/PG-1836/2020\|EPI_ISL_685442\|2020-04 | .................... | .................... | ..................... | .................... | .................... | .................... |
| V | 2088 | hCoV-19/Japan/PG-1837/2020\|EPI_ISL_685443\|2020-04 | .................... | .................... | ..................... | .................... | .................... | .................... |
| V | 2089 | hCoV-19/Japan/PG-1838/2020\|EPI_ISL_685444\|2020-04 | .................... | .................... | ..................... | .................... | .................... | .................... |
| V | 2090 | hCoV-19/Japan/PG-1839/2020\|EPI_ISL_685445\|2020-04 | .................... | .................... | ..................... | .................... | .................... | .................... |
| V | 2091 | hCoV-19/Japan/PG-1840/2020\|EPI_ISL_685446\|2020-04 | .................... | .................... | ..................... | .................... | .................... | .................... |
| V | 2092 | hCoV-19/Japan/PG-1841/2020\|EPI_ISL_685447\|2020-04 | .................... | .................... | ..................... | .................... | .................... | .................... |
| V | 2093 | hCoV-19/Japan/PG-1842/2020\|EPI_ISL_685448\|2020-04 | .................... | .................... | ..................... | .................... | .................... | .................... |
| V | 2094 | hCoV-19/Japan/PG-1843/2020\|EPI_ISL_685449\|2020-04 | .................... | .................... | ..................... | .................... | .................... | .................... |
| V | 2095 | hCoV-19/Japan/PG-1844/2020\|EPI_ISL_685450\|2020-04 | .................... | .................... | ..................... | .................... | .................... | .................... |
| V | 2096 | hCoV-19/Japan/PG-1845/2020\|EPI_ISL_685451\|2020-04 | .................... | .................... | ..................... | .................... | .................... | .................... |
| V | 2097 | hCoV-19/Japan/PG-1846/2020\|EPI_ISL_685452\|2020-04 | .................... | .................... | ..................... | .................... | .................... | .................... |
| V | 2098 | hCoV-19/Japan/PG-1847/2020\|EPI_ISL_685453\|2020-04 | .................... | .................... | ..................... | .................... | .................... | .................... |
| V | 2099 | hCoV-19/Japan/PG-1848/2020\|EPI_ISL_685454\|2020-05 | .................... | .................... | ..................... | .................... | .................... | .................... |
| V | 2100 | hCoV-19/Japan/PG-1849/2020\|EPI_ISL_685455\|2020-05 | .................... | .................... | ..................... | .................... | .................... | .................... |
| V | 2101 | hCoV-19/Japan/PG-1850/2020\|EPI_ISL_685456\|2020-05 | .................... | .................... | ..................... | .................... | .................... | .................... |
| V | 2102 | hCoV-19/Japan/PG-1851/2020\|EPI_ISL_685457\|2020-05 | .................... | .................... | ..................... | .................... | .................... | .................... |
| V | 2103 | hCoV-19/Japan/PG-1852/2020\|EPI_ISL_685458\|2020-05 | .................... | .................... | ..................... | .................... | .................... | .................... |
| V | 2104 | hCoV-19/Japan/PG-1853/2020\|EPI_ISL_685459\|2020-05 | .................... | .................... | ..................... | .................... | .................... | .................... |
| V | 2105 | hCoV-19/Japan/PG-1854/2020\|EPI_ISL_685460\|2020-05 | .................... | .................... | ..................... | .................... | .................... | .................... |
| V | 2106 | hCoV-19/Japan/PG-1855/2020\|EPI_ISL_685461\|2020-05 | .................... | .................... | ..................... | .................... | .................... | .................... |
| V | 2107 | hCoV-19/Japan/PG-1856/2020\|EPI_ISL_685462\|2020-05 | .................... | .................... | ..................... | .................... | .................... | .................... |
| V | 2108 | hCoV-19/Japan/PG-1857/2020\|EPI_ISL_685463\|2020-05 | .................... | .................... | ..................... | .................... | .................... | .................... |
| V | 2109 | hCoV-19/Japan/PG-1858/2020\|EPI_ISL_685464\|2020-05 | .................... | .................... | ..................... | .................... | .................... | .................... |
| V | 2110 | hCoV-19/Japan/PG-1859/2020\|EPI_ISL_685465\|2020-05 | .................... | .................... | ..................... | .................... | .................... | .................... |
| V | 2111 | hCoV-19/Japan/PG-1860/2020\|EPI_ISL_685466\|2020-05 | .................... | .................... | ..................... | .................... | .................... | .................... |
| V | 2112 | hCoV-19/Japan/PG-1861/2020\|EPI_ISL_685467\|2020-05 | .................... | .................... | ..................... | .................... | .................... | .................... |
| V | 2113 | hCoV-19/Japan/PG-1862/2020\|EPI_ISL_685468\|2020-05 | .................... | .................... | ..................... | .................... | .................... | .................... |
| V | 2114 | hCoV-19/Japan/PG-1863/2020\|EPI_ISL_685469\|2020-05 | .................... | .................... | ..................... | .................... | .................... | .................... |
| V | 2115 | hCoV-19/Japan/PG-1864/2020\|EPI_ISL_685470\|2020-05 | .................... | .................... | ..................... | .................... | .................... | .................... |
| V | 2116 | hCoV-19/Japan/PG-1865/2020\|EPI_ISL_685471\|2020-05 | .................... | .................... | ..................... | .................... | .................... | .................... |
| V | 2117 | hCoV-19/Japan/PG-1866/2020\|EPI_ISL_685472\|2020-05 | .................... | .................... | ..................... | .................... | .................... | .................... |
| V | 2118 | hCoV-19/Japan/PG-1867/2020\|EPI_ISL_685473\|2020-05 | .................... | .................... | ..................... | .................... | .................... | .................... |
| V | 2119 | hCoV-19/Japan/PG-1868/2020\|EPI_ISL_685474\|2020-05 | .................... | .................... | ..................... | .................... | .................... | .................... |
| V | 2120 | hCoV-19/Japan/PG-1869/2020\|EPI_ISL_685475\|2020-05 | .................... | .................... | ..................... | .................... | .................... | .................... |
| V | 2121 | hCoV-19/Japan/PG-1870/2020\|EPI_ISL_685476\|2020-05 | .................... | .................... | ..................... | .................... | .................... | .................... |
| V | 2122 | hCoV-19/Japan/PG-1871/2020\|EPI_ISL_685477\|2020-05 | .................... | .................... | ..................... | .................... | .................... | .................... |
| V | 2123 | hCoV-19/Japan/PG-1872/2020\|EPI_ISL_685478\|2020-05 | .................... | .................... | ..................... | .................... | .................... | .................... |
| V | 2124 | hCoV-19/Japan/PG-1873/2020\|EPI_ISL_685479\|2020-05 | .................... | .................... | ..................... | .................... | .................... | .................... |
| V | 2125 | hCoV-19/Japan/PG-1874/2020\|EPI_ISL_685480\|2020-05 | .................... | .................... | ..................... | .................... | .................... | .................... |
| V | 2126 | hCoV-19/Japan/PG-1875/2020\|EPI_ISL_685481\|2020-05 | .................... | .................... | ..................... | .................... | .................... | .................... |
| V | 2127 | hCoV-19/Japan/PG-1876/2020\|EPI_ISL_685482\|2020-05 | .................... | .................... | ..................... | .................... | .................... | .................... |
| V | 2128 | hCoV-19/Japan/PG-1877/2020\|EPI_ISL_685483\|2020-05 | .................... | .................... | ..................... | .................... | .................... | .................... |
| V | 2129 | hCoV-19/Japan/PG-1878/2020\|EPI_ISL_685484\|2020-05 | .................... | .................... | ..................... | .................... | .................... | .................... |
| V | 2130 | hCoV-19/Japan/PG-1879/2020\|EPI_ISL_685485\|2020-05 | .................... | .................... | ..................... | .................... | .................... | .................... |
| V | 2131 | hCoV-19/Japan/PG-1880/2020\|EPI_ISL_685486\|2020-05 | .................... | .................... | ..................... | .................... | .................... | .................... |
| V | 2132 | hCoV-19/Japan/PG-1881/2020\|EPI_ISL_685487\|2020-05 | .................... | .................... | ..................... | .................... | .................... | .................... |
| V | 2133 | hCoV-19/Japan/PG-1882/2020\|EPI_ISL_685488\|2020-05 | .................... | .................... | ..................... | .................... | .................... | .................... |
| V | 2134 | hCoV-19/Japan/PG-7598/2020\|EPI_ISL_686700\|2020-07 | .................... | .................... | ..................... | .................... | .................... | .................... |
| V | 2135 | hCoV-19/Japan/PG-7599/2020\|EPI_ISL_686701\|2020-07 | .................... | .................... | ..................... | .................... | .................... | .................... |

V 2136 hCoV-19/Japan/PG-7600/2020|EPI_ISL_686702|2020-07 .................... .................... ..................... .................... .................... ....................
V 2137 hCoV-19/Japan/PG-7601/2020|EPI_ISL_686703|2020-07 .................... .................... ..................... .................... .................... ....................
V 2138 hCoV-19/Japan/PG-7602/2020|EPI_ISL_686704|2020-07 .................... .................... ..................... .................... .................... ....................
V 2139 hCoV-19/Japan/PG-7603/2020|EPI_ISL_686705|2020-07 .................... .................... ..................... .................... .................... ....................
V 2140 hCoV-19/Japan/PG-7604/2020|EPI_ISL_686706|2020-07 .................... .................... ..................... .................... .................... ....................
V 2141 hCoV-19/Japan/PG-7605/2020|EPI_ISL_686707|2020-07 .................... .................... ..................... .................... .................... ....................
V 2142 hCoV-19/Japan/PG-7606/2020|EPI_ISL_686708|2020-07 .................... .................... ..................... .................... .................... ....................
V 2143 hCoV-19/Japan/PG-7607/2020|EPI_ISL_686709|2020-07 .................... .................... ..................... .................... .................... ....................
V 2144 hCoV-19/Japan/PG-7608/2020|EPI_ISL_686710|2020-07 .................... .................... ..................... .................... .................... ....................
V 2145 hCoV-19/Japan/PG-7609/2020|EPI_ISL_686711|2020-07 .................... .................... ..................... .................... .................... ....................
V 2146 hCoV-19/Japan/PG-7610/2020|EPI_ISL_686712|2020-07 .................... .................... ..................... .................... .................... ....................
V 2147 hCoV-19/Japan/PG-7611/2020|EPI_ISL_686713|2020-07 .................... .................... ..................... .................... .................... ....................
V 2148 hCoV-19/Japan/PG-7612/2020|EPI_ISL_686714|2020-07 .................... .................... ..................... .................... .................... ....................
V 2149 hCoV-19/Japan/PG-7613/2020|EPI_ISL_686715|2020-07 .................... .................... ..................... .................... .................... ....................
V 2150 hCoV-19/Japan/PG-7614/2020|EPI_ISL_686716|2020-07 .................... .................... ..................... .................... .................... ....................
V 2151 hCoV-19/Japan/PG-7615/2020|EPI_ISL_686717|2020-08 .................... .................... ..................... .................... .................... ....................
V 2152 hCoV-19/Japan/PG-7616/2020|EPI_ISL_686718|2020-08 .................... .................... ..................... .................... .................... ....................
V 2153 hCoV-19/Japan/PG-7617/2020|EPI_ISL_686719|2020-08 .................... .................... ..................... .................... .................... ....................
V 2154 hCoV-19/Japan/PG-7618/2020|EPI_ISL_686720|2020-08 .................... .................... ..................... .................... .................... ....................
V 2155 hCoV-19/Japan/PG-7619/2020|EPI_ISL_686721|2020-08 .................... .................... ..................... .................... .................... ....................
V 2156 hCoV-19/Japan/PG-7620/2020|EPI_ISL_686722|2020-08 .................... .................... ..................... .................... .................... ....................
V 2157 hCoV-19/Japan/PG-7621/2020|EPI_ISL_686723|2020-08 .................... .................... ..................... .................... .................... ....................
V 2158 hCoV-19/Japan/PG-7622/2020|EPI_ISL_686724|2020-08 .................... .................... ..................... .................... .................... ....................
V 2159 hCoV-19/Japan/PG-7623/2020|EPI_ISL_686725|2020-08 .................... .................... ..................... .................... .................... ....................
V 2160 hCoV-19/Japan/PG-7624/2020|EPI_ISL_686726|2020-08 .................... .................... ..................... .................... .................... ....................
V 2161 hCoV-19/Japan/PG-7625/2020|EPI_ISL_686727|2020-08 .................... .................... ..................... .................... .................... ....................
V 2162 hCoV-19/Japan/PG-7626/2020|EPI_ISL_686728|2020-08 .................... .................... ..................... .................... .................... ....................
V 2163 hCoV-19/Japan/PG-7627/2020|EPI_ISL_686729|2020-08 .................... .................... ..................... .................... .................... ....................
V 2164 hCoV-19/Japan/PG-7628/2020|EPI_ISL_686730|2020-08 .................... .................... ..................... .................... .................... ....................
V 2165 hCoV-19/Japan/PG-7629/2020|EPI_ISL_686731|2020-08 .................... .................... ..................... .................... .................... ....................
V 2166 hCoV-19/Japan/PG-7668/2020|EPI_ISL_686770|2020-08 .................... .................... ..................... .................... .................... ....................
V 2167 hCoV-19/Japan/PG-7669/2020|EPI_ISL_686771|2020-08 .................... .................... ..................... .................... .................... ....................
V 2168 hCoV-19/Japan/PG-7670/2020|EPI_ISL_686772|2020-08 .................... .................... ..................... .................... .................... ....................
V 2169 hCoV-19/Japan/PG-7671/2020|EPI_ISL_686773|2020-08 .................... .................... ..................... .................... .................... ....................
V 2170 hCoV-19/Japan/PG-7672/2020|EPI_ISL_686774|2020-08 .................... .................... ..................... .................... .................... ....................
V 2171 hCoV-19/Japan/PG-7673/2020|EPI_ISL_686775|2020-08 .................... .................... ..................... .................... .................... ....................
V 2172 hCoV-19/Japan/PG-7674/2020|EPI_ISL_686776|2020-08 .................... .................... ..................... .................... .................... ....................
V 2173 hCoV-19/Japan/PG-7675/2020|EPI_ISL_686777|2020-08 .................... .................... ..................... .................... .................... ....................
V 2174 hCoV-19/Japan/PG-7676/2020|EPI_ISL_686778|2020-08 .................... .................... ..................... .................... .................... ....................
V 2175 hCoV-19/Japan/PG-7677/2020|EPI_ISL_686779|2020-08 .................... .................... ..................... .................... .................... ....................
V 2176 hCoV-19/Japan/PG-7678/2020|EPI_ISL_686780|2020-08 .................... .................... ..................... .................... .................... ....................
V 2177 hCoV-19/Japan/PG-7679/2020|EPI_ISL_686781|2020-08 .................... .................... ..................... .................... .................... ....................
V 2178 hCoV-19/Japan/PG-7680/2020|EPI_ISL_686782|2020-08 .................... .................... ..................... .................... .................... ....................
V 2179 hCoV-19/Japan/PG-7681/2020|EPI_ISL_686783|2020-08 .................... .................... ..................... .................... .................... ....................
V 2180 hCoV-19/Japan/PG-7682/2020|EPI_ISL_686784|2020-08 .................... .................... ..................... .................... .................... ....................
V 2181 hCoV-19/Japan/PG-7683/2020|EPI_ISL_686785|2020-08 .................... .................... ..................... .................... .................... ....................
V 2182 hCoV-19/Japan/PG-7684/2020|EPI_ISL_686786|2020-08 .................... .................... ..................... .................... .................... ....................
V 2183 hCoV-19/Japan/PG-7685/2020|EPI_ISL_686787|2020-08 .................... .................... ..................... .................... .................... ....................
V 2184 hCoV-19/Japan/PG-7686/2020|EPI_ISL_686788|2020-08 .................... .................... ..................... .................... .................... ....................
V 2185 hCoV-19/Japan/PG-7687/2020|EPI_ISL_686789|2020-08 .................... .................... ..................... .................... .................... ....................
V 2186 hCoV-19/Japan/PG-7688/2020|EPI_ISL_686790|2020-08 .................... .................... ..................... .................... .................... ....................
V 2187 hCoV-19/Japan/PG-7689/2020|EPI_ISL_686791|2020-08 .................... .................... ..................... .................... .................... ....................
V 2188 hCoV-19/Japan/PG-7690/2020|EPI_ISL_686792|2020-08 .................... .................... ..................... .................... .................... ....................
V 2189 hCoV-19/Japan/PG-7691/2020|EPI_ISL_686793|2020-08 .................... .................... ..................... .................... .................... ....................
V 2190 hCoV-19/Japan/PG-7692/2020|EPI_ISL_686794|2020-08 .................... .................... ..................... .................... .................... ....................
V 2191 hCoV-19/Japan/PG-7693/2020|EPI_ISL_686795|2020-08 .................... .................... ..................... .................... .................... ....................
V 2192 hCoV-19/Japan/PG-7694/2020|EPI_ISL_686796|2020-08 .................... .................... ..................... .................... .................... ....................
V 2193 hCoV-19/Japan/PG-7695/2020|EPI_ISL_686797|2020-08 .................... .................... ..................... .................... .................... ....................
V 2194 hCoV-19/Japan/PG-7696/2020|EPI_ISL_686798|2020-08 .................... .................... ..................... .................... .................... ....................
V 2195 hCoV-19/Japan/PG-7697/2020|EPI_ISL_686799|2020-08 .................... .................... ..................... .................... .................... ....................
V 2196 hCoV-19/Japan/PG-3120/2020|EPI_ISL_691242|2020-03 .................... .................... ..................... .................... .................... ....................
V 2197 hCoV-19/Japan/PG-3121/2020|EPI_ISL_691243|2020-04 .................... .................... ..................... .................... .................... ....................
V 2198 hCoV-19/Japan/PG-3122/2020|EPI_ISL_691244|2020-04 .................... .................... ..................... .................... .................... ....................
V 2199 hCoV-19/Japan/PG-3123/2020|EPI_ISL_691245|2020-04 .................... .................... ..................... .................... .................... ....................
V 2200 hCoV-19/Japan/PG-3124/2020|EPI_ISL_691246|2020-04 .................... .................... ..................... .................... .................... ....................
V 2201 hCoV-19/Japan/PG-3125/2020|EPI_ISL_691247|2020-04 .................... .................... ..................... .................... .................... ....................
V 2202 hCoV-19/Japan/PG-3126/2020|EPI_ISL_691248|2020-05 .................... .................... ..................... .................... .................... ....................
V 2203 hCoV-19/Japan/PG-3127/2020|EPI_ISL_691249|2020-04 .................... .................... ..................... .................... .................... ....................
V 2204 hCoV-19/Japan/PG-3128/2020|EPI_ISL_691250|2020-04 .................... .................... ..................... .................... .................... ....................
V 2205 hCoV-19/Japan/PG-3129/2020|EPI_ISL_691251|2020-04 .................... .................... ..................... .................... .................... ....................
V 2206 hCoV-19/Japan/PG-3130/2020|EPI_ISL_691252|2020-06 .................... .................... ..................... .................... .................... ....................
V 2207 hCoV-19/Japan/PG-3131/2020|EPI_ISL_691253|2020-06 .................... .................... ..................... .................... .................... ....................
V 2208 hCoV-19/Japan/PG-3132/2020|EPI_ISL_691254|2020-06 .................... .................... ..................... .................... .................... ....................
V 2209 hCoV-19/Japan/PG-3133/2020|EPI_ISL_691255|2020-06 .................... .................... ..................... .................... .................... ....................
V 2210 hCoV-19/Japan/PG-3134/2020|EPI_ISL_691256|2020-06 .................... .................... ..................... .................... .................... ....................
V 2211 hCoV-19/Japan/PG-3135/2020|EPI_ISL_691257|2020-04 .................... .................... ..................... .................... .................... ....................
V 2212 hCoV-19/Japan/PG-3136/2020|EPI_ISL_691258|2020-04 .................... .................... ..................... .................... .................... ....................
V 2213 hCoV-19/Japan/PG-3137/2020|EPI_ISL_691259|2020-05 .................... .................... ..................... .................... .................... ....................
V 2214 hCoV-19/Japan/PG-3138/2020|EPI_ISL_691260|2020-06 .................... .................... ..................... .................... .................... ....................
V 2215 hCoV-19/Japan/PG-3139/2020|EPI_ISL_691261|2020-06 .................... .................... ..................... .................... .................... ....................

| | | | | | | | | |
|---|---|---|---|---|---|---|---|---|
| V | 2216 | hCoV-19/Japan/PG-3140/2020\|EPI_ISL_691262\|2020-06 | ···················· | ···················· | ····················· | ···················· | ···················· | ···················· |
| V | 2217 | hCoV-19/Japan/PG-3141/2020\|EPI_ISL_691263\|2020-06 | ···················· | ···················· | ····················· | ···················· | ···················· | ···················· |
| V | 2218 | hCoV-19/Japan/PG-3142/2020\|EPI_ISL_691264\|2020-06 | ···················· | ···················· | ····················· | ···················· | ···················· | ···················· |
| V | 2219 | hCoV-19/Japan/PG-3143/2020\|EPI_ISL_691265\|2020-06 | ···················· | ···················· | ····················· | ···················· | ···················· | ···················· |
| V | 2220 | hCoV-19/Japan/PG-3144/2020\|EPI_ISL_691266\|2020-06 | ···················· | ···················· | ····················· | ···················· | ···················· | ···················· |
| V | 2221 | hCoV-19/Japan/PG-3145/2020\|EPI_ISL_691267\|2020-06 | ···················· | ···················· | ····················· | ···················· | ···················· | ···················· |
| V | 2222 | hCoV-19/Japan/PG-3147/2020\|EPI_ISL_691268\|2020-06 | ···················· | ···················· | ····················· | ···················· | ···················· | ···················· |
| V | 2223 | hCoV-19/Japan/PG-3148/2020\|EPI_ISL_691269\|2020-06 | ···················· | ···················· | ····················· | ···················· | ···················· | ···················· |
| V | 2224 | hCoV-19/Japan/PG-3150/2020\|EPI_ISL_691270\|2020-06 | ···················· | ···················· | ····················· | ···················· | ···················· | ···················· |
| V | 2225 | hCoV-19/Japan/PG-3151/2020\|EPI_ISL_691271\|2020-06 | ···················· | ···················· | ····················· | ···················· | ···················· | ···················· |
| V | 2226 | hCoV-19/Japan/PG-3152/2020\|EPI_ISL_691272\|2020-06 | ···················· | ···················· | ····················· | ···················· | ···················· | ···················· |
| V | 2227 | hCoV-19/Japan/PG-3153/2020\|EPI_ISL_691273\|2020-05 | ···················· | ···················· | ····················· | ···················· | ···················· | ···················· |
| V | 2228 | hCoV-19/Japan/PG-3154/2020\|EPI_ISL_691274\|2020-05 | ···················· | ···················· | ····················· | ···················· | ···················· | ···················· |
| V | 2229 | hCoV-19/Japan/PG-3155/2020\|EPI_ISL_691275\|2020-06 | ···················· | ···················· | ····················· | ···················· | ···················· | ···················· |
| V | 2230 | hCoV-19/Japan/PG-3156/2020\|EPI_ISL_691276\|2020-06 | ···················· | ···················· | ····················· | ···················· | ···················· | ···················· |
| V | 2231 | hCoV-19/Japan/PG-3157/2020\|EPI_ISL_691277\|2020-06 | ···················· | ···················· | ····················· | ···················· | ···················· | ···················· |
| V | 2232 | hCoV-19/Japan/PG-3158/2020\|EPI_ISL_691278\|2020-06 | ···················· | ···················· | ····················· | ···················· | ···················· | ···················· |
| V | 2233 | hCoV-19/Japan/PG-3159/2020\|EPI_ISL_691279\|2020-06 | ···················· | ···················· | ····················· | ···················· | ···················· | ···················· |
| V | 2234 | hCoV-19/Japan/PG-3160/2020\|EPI_ISL_691280\|2020-06 | ···················· | ···················· | ····················· | ···················· | ···················· | ···················· |
| V | 2235 | hCoV-19/Japan/PG-3161/2020\|EPI_ISL_691281\|2020-06 | ···················· | ···················· | ····················· | ···················· | ···················· | ···················· |
| V | 2236 | hCoV-19/Japan/PG-3162/2020\|EPI_ISL_691282\|2020-06 | ···················· | ···················· | ····················· | ···················· | ···················· | ···················· |
| V | 2237 | hCoV-19/Japan/PG-3163/2020\|EPI_ISL_691283\|2020-06 | ···················· | ···················· | ····················· | ···················· | ···················· | ···················· |
| V | 2238 | hCoV-19/Japan/PG-3164/2020\|EPI_ISL_691284\|2020-06 | ···················· | ···················· | ····················· | ···················· | ···················· | ···················· |
| V | 2239 | hCoV-19/Japan/PG-3165/2020\|EPI_ISL_691285\|2020-06 | ···················· | ···················· | ····················· | ···················· | ···················· | ···················· |
| V | 2240 | hCoV-19/Japan/PG-3166/2020\|EPI_ISL_691286\|2020-06 | ···················· | ···················· | ····················· | ···················· | ···················· | ···················· |
| V | 2241 | hCoV-19/Japan/PG-3167/2020\|EPI_ISL_691287\|2020-06 | ···················· | ···················· | ····················· | ···················· | ···················· | ···················· |
| V | 2242 | hCoV-19/Japan/PG-3168/2020\|EPI_ISL_691288\|2020-06 | ···················· | ···················· | ····················· | ···················· | ···················· | ···················· |
| V | 2243 | hCoV-19/Japan/PG-3169/2020\|EPI_ISL_691289\|2020-03 | ···················· | ···················· | ····················· | ···················· | ···················· | ···················· |
| V | 2244 | hCoV-19/Japan/PG-3171/2020\|EPI_ISL_691291\|2020-03 | ···················· | ···················· | ····················· | ···················· | ···················· | ···················· |
| V | 2245 | hCoV-19/Japan/PG-4345/2020\|EPI_ISL_692573\|2020-07 | ···················· | ···················· | ····················· | ···················· | ···················· | ···················· |
| V | 2246 | hCoV-19/Japan/PG-4346/2020\|EPI_ISL_692574\|2020-07 | ···················· | ···················· | ····················· | ···················· | ···················· | ···················· |
| V | 2247 | hCoV-19/Japan/PG-4347/2020\|EPI_ISL_692575\|2020-07 | ···················· | ···················· | ····················· | ···················· | ···················· | ···················· |
| V | 2248 | hCoV-19/Japan/PG-4348/2020\|EPI_ISL_692576\|2020-07 | ···················· | ···················· | ····················· | ···················· | ···················· | ···················· |
| V | 2249 | hCoV-19/Japan/PG-4349/2020\|EPI_ISL_692577\|2020-07 | ···················· | ···················· | ····················· | ···················· | ···················· | ···················· |
| V | 2250 | hCoV-19/Japan/PG-4350/2020\|EPI_ISL_692578\|2020-07 | ···················· | ···················· | ····················· | ···················· | ···················· | ···················· |
| V | 2251 | hCoV-19/Japan/PG-4351/2020\|EPI_ISL_692579\|2020-07 | ···················· | ···················· | ····················· | ···················· | ···················· | ···················· |
| V | 2252 | hCoV-19/Japan/PG-4352/2020\|EPI_ISL_692580\|2020-07 | ···················· | ···················· | ····················· | ···················· | ···················· | ···················· |
| V | 2253 | hCoV-19/Japan/PG-4353/2020\|EPI_ISL_692581\|2020-07 | ···················· | ···················· | ····················· | ···················· | ···················· | ···················· |
| V | 2254 | hCoV-19/Japan/PG-4354/2020\|EPI_ISL_692582\|2020-07 | ···················· | ···················· | ····················· | ···················· | ···················· | ···················· |
| V | 2255 | hCoV-19/Japan/PG-4355/2020\|EPI_ISL_692583\|2020-07 | ···················· | ···················· | ····················· | ···················· | ···················· | ···················· |
| V | 2256 | hCoV-19/Japan/PG-4356/2020\|EPI_ISL_692584\|2020-07 | ···················· | ···················· | ····················· | ···················· | ···················· | ···················· |
| V | 2257 | hCoV-19/Japan/PG-4357/2020\|EPI_ISL_692585\|2020-07 | ···················· | ···················· | ····················· | ···················· | ···················· | ···················· |
| V | 2258 | hCoV-19/Japan/PG-4358/2020\|EPI_ISL_692586\|2020-07 | ···················· | ·A·················· | ····················· | ···················· | ···················· | ···················· |
| V | 2259 | hCoV-19/Japan/PG-4359/2020\|EPI_ISL_692587\|2020-07 | ···················· | ···················· | ····················· | ···················· | ···················· | ···················· |
| V | 2260 | hCoV-19/Japan/PG-4360/2020\|EPI_ISL_692588\|2020-07 | ···················· | ···················· | ····················· | ···················· | ···················· | ···················· |
| V | 2261 | hCoV-19/Japan/PG-4361/2020\|EPI_ISL_692589\|2020-07 | ···················· | ···················· | ····················· | ···················· | ···················· | ···················· |
| V | 2262 | hCoV-19/Japan/PG-4362/2020\|EPI_ISL_692590\|2020-07 | ···················· | ···················· | ····················· | ···················· | ···················· | ···················· |
| V | 2263 | hCoV-19/Japan/PG-4363/2020\|EPI_ISL_692591\|2020-07 | ···················· | ···················· | ····················· | ···················· | ···················· | ···················· |
| V | 2264 | hCoV-19/Japan/PG-4365/2020\|EPI_ISL_692592\|2020-07 | ···················· | ···················· | ····················· | ···················· | ···················· | ···················· |
| V | 2265 | hCoV-19/Japan/PG-4366/2020\|EPI_ISL_692593\|2020-07 | ···················· | ···················· | ····················· | ···················· | ···················· | ···················· |
| V | 2266 | hCoV-19/Japan/PG-4367/2020\|EPI_ISL_692594\|2020-07 | ···················· | ···················· | ····················· | ···················· | ···················· | ···················· |
| V | 2267 | hCoV-19/Japan/PG-4368/2020\|EPI_ISL_692595\|2020-07 | ···················· | ···················· | ····················· | ···················· | ···················· | ···················· |
| V | 2268 | hCoV-19/Japan/PG-4369/2020\|EPI_ISL_692596\|2020-07 | ···················· | ···················· | ····················· | ···················· | ···················· | ···················· |
| V | 2269 | hCoV-19/Japan/PG-4370/2020\|EPI_ISL_692597\|2020-05 | ···················· | ···················· | ····················· | ···················· | ···················· | ···················· |
| V | 2270 | hCoV-19/Japan/PG-4371/2020\|EPI_ISL_692598\|2020-05 | ···················· | ···················· | ····················· | ···················· | ···················· | ···················· |
| V | 2271 | hCoV-19/Japan/PG-4372/2020\|EPI_ISL_692599\|2020-05 | ···················· | ···················· | ····················· | ···················· | ···················· | ···················· |
| V | 2272 | hCoV-19/Japan/PG-4373/2020\|EPI_ISL_692600\|2020-06 | ···················· | ···················· | ····················· | ···················· | ···················· | ···················· |
| V | 2273 | hCoV-19/Japan/PG-4374/2020\|EPI_ISL_692601\|2020-06 | ···················· | ···················· | ····················· | ···················· | ···················· | ···················· |
| V | 2274 | hCoV-19/Japan/PG-4375/2020\|EPI_ISL_692602\|2020-06 | ···················· | ···················· | ····················· | ···················· | ···················· | ···················· |
| V | 2275 | hCoV-19/Japan/PG-4376/2020\|EPI_ISL_692603\|2020-06 | ···················· | ···················· | ····················· | ···················· | ···················· | ···················· |
| V | 2276 | hCoV-19/Japan/PG-4377/2020\|EPI_ISL_692604\|2020-06 | ···················· | ···················· | ····················· | ···················· | ···················· | ···················· |
| V | 2277 | hCoV-19/Japan/PG-4378/2020\|EPI_ISL_692605\|2020-06 | ···················· | ···················· | ····················· | ···················· | ···················· | ···················· |
| V | 2278 | hCoV-19/Japan/PG-4379/2020\|EPI_ISL_692606\|2020-06 | ···················· | ···················· | ····················· | ···················· | ···················· | ···················· |
| V | 2279 | hCoV-19/Japan/PG-4380/2020\|EPI_ISL_692607\|2020-06 | ···················· | ···················· | ····················· | ···················· | ···················· | ···················· |
| V | 2280 | hCoV-19/Japan/PG-4381/2020\|EPI_ISL_692608\|2020-06 | ···················· | ···················· | ····················· | ···················· | ···················· | ···················· |
| V | 2281 | hCoV-19/Japan/PG-4382/2020\|EPI_ISL_692609\|2020-06 | ···················· | ···················· | ····················· | ···················· | ···················· | ···················· |
| V | 2282 | hCoV-19/Switzerland/SZ-ETHZ-360105/2020\|EPI_ISL_69 | ···················· | ···················· | ····················· | ···················· | ···················· | ···················· |
| V | 2283 | hCoV-19/Switzerland/ZH-ETHZ-360116/2020\|EPI_ISL_69 | ···················· | ···················· | ····················· | ···················· | ···················· | ···················· |
| V | 2284 | hCoV-19/Switzerland/GE-ETHZ-360137/2020\|EPI_ISL_69 | ···················· | ···················· | ····················· | ···················· | ···················· | ···················· |
| V | 2285 | hCoV-19/Switzerland/BL-ETHZ-360148/2020\|EPI_ISL_69 | ···················· | ···················· | ····················· | ···················· | ···················· | ···················· |
| V | 2286 | hCoV-19/Switzerland/BL-ETHZ-360149/2020\|EPI_ISL_69 | ···················· | ···················· | ····················· | ···················· | ···················· | ···················· |
| V | 2287 | hCoV-19/Switzerland/BL-ETHZ-360150/2020\|EPI_ISL_69 | ···················· | ···················· | ····················· | ···················· | ···················· | ···················· |
| V | 2288 | hCoV-19/Switzerland/BL-ETHZ-360151/2020\|EPI_ISL_69 | ···················· | ···················· | ····················· | ···················· | ···················· | ···················· |
| V | 2289 | hCoV-19/Switzerland/BL-ETHZ-360152/2020\|EPI_ISL_69 | ···················· | ···················· | ····················· | ···················· | ···················· | ···················· |
| V | 2290 | hCoV-19/Switzerland/BL-ETHZ-360153/2020\|EPI_ISL_69 | ···················· | ·A·················· | ····················· | ···················· | ···················· | ··A················· |
| V | 2291 | hCoV-19/Switzerland/BL-ETHZ-360154/2020\|EPI_ISL_69 | ···················· | ···················· | ····················· | ···················· | ···················· | ···················· |
| V | 2292 | hCoV-19/Switzerland/BL-ETHZ-360155/2020\|EPI_ISL_69 | ···················· | ·A·················· | ····················· | ···················· | ···················· | ··A················· |
| V | 2293 | hCoV-19/Switzerland/BL-ETHZ-360156/2020\|EPI_ISL_69 | ···················· | ···················· | ····················· | ···················· | ···················· | ···················· |
| V | 2294 | hCoV-19/Switzerland/SO-ETHZ-360158/2020\|EPI_ISL_69 | ···················· | ···················· | ····················· | ···················· | ···················· | ···················· |
| V | 2295 | hCoV-19/Switzerland/SO-ETHZ-360159/2020\|EPI_ISL_69 | ···················· | ···················· | ····················· | ···················· | ···················· | ···················· |

V 2296 hCoV-19/Switzerland/SO-ETHZ-360160/2020|EPI_ISL_69 .................... .................... .................... .................... .................... ....................
V 2297 hCoV-19/Switzerland/BL-ETHZ-360161/2020|EPI_ISL_69 .................... .................... .................... .................... .................... ....................
V 2298 hCoV-19/Switzerland/SO-ETHZ-360165/2020|EPI_ISL_69 .................... .................... .................... .................... .................... ....................
V 2299 hCoV-19/Switzerland/SO-ETHZ-360166/2020|EPI_ISL_69 .................... .................... .................... .................... .................... ....................
V 2300 hCoV-19/Switzerland/SO-ETHZ-360167/2020|EPI_ISL_69 .................... .................... .................... .................... .................... ....................
V 2301 hCoV-19/Switzerland/SO-ETHZ-360168/2020|EPI_ISL_69 .................... .................... .................... .................... .................... ....................
V 2302 hCoV-19/Switzerland/SO-ETHZ-360169/2020|EPI_ISL_69 .................... .................... .................... .................... .................... ....................
V 2303 hCoV-19/Switzerland/SO-ETHZ-360170/2020|EPI_ISL_69 .................... .................... .................... .................... .................... ....................
V 2304 hCoV-19/Switzerland/BS-ETHZ-370264/2020|EPI_ISL_69 .................... .................... .................... .................... .................... ....................
V 2305 hCoV-19/France/ARA-29628/2020|EPI_ISL_684107|2020- .................... .................... .................... .................... .................... ....................
V 2306 hCoV-19/Japan/PG-1774/2020|EPI_ISL_685380|2020-04 .................... .................... .................... .................... .................... ....................
V 2307 hCoV-19/Japan/PG-1775/2020|EPI_ISL_685381|2020-04 .................... .................... .................... .................... .................... ....................
V 2308 hCoV-19/Japan/PG-1776/2020|EPI_ISL_685382|2020-04 .................... .................... .................... .................... .................... ....................
V 2309 hCoV-19/Japan/PG-1777/2020|EPI_ISL_685383|2020-04 .................... .................... .................... .................... .................... ....................
V 2310 hCoV-19/Japan/PG-1778/2020|EPI_ISL_685384|2020-04 .................... .................... .................... .................... .................... ....................
V 2311 hCoV-19/Japan/PG-1779/2020|EPI_ISL_685385|2020-04 .................... .................... .................... .................... .................... ....................
V 2312 hCoV-19/Japan/PG-1780/2020|EPI_ISL_685386|2020-04 .................... .................... .................... .................... .................... ....................
V 2313 hCoV-19/Japan/PG-1781/2020|EPI_ISL_685387|2020-04 .................... .................... .................... .................... .................... ....................
V 2314 hCoV-19/Japan/PG-1782/2020|EPI_ISL_685388|2020-04 .................... .................... .................... .................... .................... ....................
V 2315 hCoV-19/Japan/PG-1783/2020|EPI_ISL_685389|2020-04 .................... .................... .................... .................... .................... ....................
V 2316 hCoV-19/Japan/PG-1784/2020|EPI_ISL_685390|2020-05 .................... .................... .................... .................... .................... ....................
V 2317 hCoV-19/Japan/PG-1785/2020|EPI_ISL_685391|2020-05 .................... .................... .................... .................... .................... ....................
V 2318 hCoV-19/Japan/PG-1786/2020|EPI_ISL_685392|2020-05 .................... .................... .................... .................... .................... ....................
V 2319 hCoV-19/Japan/PG-1787/2020|EPI_ISL_685393|2020-05 .................... .................... .................... .................... .................... ....................
V 2320 hCoV-19/Japan/PG-1788/2020|EPI_ISL_685394|2020-05 .................... .................... .................... .................... .................... ....................
V 2321 hCoV-19/Japan/PG-1789/2020|EPI_ISL_685395|2020-05 .................... .................... .................... .................... .................... ....................
V 2322 hCoV-19/Japan/PG-1790/2020|EPI_ISL_685396|2020-05 .................... .................... .................... .................... .................... ....................
V 2323 hCoV-19/Japan/PG-1791/2020|EPI_ISL_685397|2020-05 .................... .................... .................... .................... .................... ....................
V 2324 hCoV-19/Japan/PG-1795/2020|EPI_ISL_685401|2020-05 .................... .................... .................... .................... .................... ....................
V 2325 hCoV-19/Japan/PG-1796/2020|EPI_ISL_685402|2020-05 .................... .................... .................... .................... .................... ....................
V 2326 hCoV-19/Japan/PG-1797/2020|EPI_ISL_685403|2020-05 .................... .................... .................... .................... .................... ....................
V 2327 hCoV-19/Japan/PG-1798/2020|EPI_ISL_685404|2020-05 .................... .................... .................... .................... .................... ....................
V 2328 hCoV-19/Japan/PG-1799/2020|EPI_ISL_685405|2020-05 .................... .................... .................... .................... .................... ....................
V 2329 hCoV-19/Japan/PG-1800/2020|EPI_ISL_685406|2020-05 .................... .................... .................... .................... .................... ....................
V 2330 hCoV-19/Japan/PG-1801/2020|EPI_ISL_685407|2020-04 .................... .................... .................... .................... .................... ....................
V 2331 hCoV-19/Japan/PG-1802/2020|EPI_ISL_685408|2020-04 .................... .................... .................... .................... .................... ....................
V 2332 hCoV-19/Japan/PG-1803/2020|EPI_ISL_685409|2020-04 .................... .................... .................... .................... .................... ....................
V 2333 hCoV-19/Japan/PG-1804/2020|EPI_ISL_685410|2020-04 .................... .................... .................... .................... .................... ....................
V 2334 hCoV-19/Japan/PG-1805/2020|EPI_ISL_685411|2020-04 .................... .................... .................... .................... .................... ....................
V 2335 hCoV-19/Japan/PG-1806/2020|EPI_ISL_685412|2020-04 .................... .................... .................... .................... .................... ....................
V 2336 hCoV-19/Japan/PG-1807/2020|EPI_ISL_685413|2020-04 .................... .................... .................... .................... .................... ....................
V 2337 hCoV-19/Japan/PG-1808/2020|EPI_ISL_685414|2020-04 .................... .................... .................... .................... .................... ....................
V 2338 hCoV-19/Japan/PG-1809/2020|EPI_ISL_685415|2020-04 .................... .................... .................... .................... .................... ....................
V 2339 hCoV-19/Japan/PG-1810/2020|EPI_ISL_685416|2020-04 .................... .................... .................... .................... .................... ....................
V 2340 hCoV-19/Japan/PG-1811/2020|EPI_ISL_685417|2020-04 .................... .................... .................... .................... .................... ....................
V 2341 hCoV-19/Japan/PG-1812/2020|EPI_ISL_685418|2020-04 .................... .................... .................... .................... .................... ....................
V 2342 hCoV-19/Japan/PG-1813/2020|EPI_ISL_685419|2020-04 .................... .................... .................... .................... .................... ....................
V 2343 hCoV-19/Japan/PG-1814/2020|EPI_ISL_685420|2020-04 .................... .................... .................... .................... .................... ....................
V 2344 hCoV-19/Japan/PG-1815/2020|EPI_ISL_685421|2020-04 .................... .................... .................... .................... .................... ....................
V 2345 hCoV-19/Japan/PG-1816/2020|EPI_ISL_685422|2020-04 .................... .................... .................... .................... .................... ....................
V 2346 hCoV-19/Japan/PG-1817/2020|EPI_ISL_685423|2020-04 .................... .................... .................... .................... .................... ....................
V 2347 hCoV-19/Japan/PG-1818/2020|EPI_ISL_685424|2020-04 .................... .................... .................... .................... .................... ....................
V 2348 hCoV-19/Japan/PG-1819/2020|EPI_ISL_685425|2020-04 .................... .................... .................... .................... .................... ....................
V 2349 hCoV-19/Japan/PG-1820/2020|EPI_ISL_685426|2020-04 .................... .................... .................... .................... .................... ....................
V 2350 hCoV-19/Japan/PG-1821/2020|EPI_ISL_685427|2020-04 .................... .................... .................... .................... .................... ....................
V 2351 hCoV-19/Japan/PG-1822/2020|EPI_ISL_685428|2020-04 .................... .................... .................... .................... .................... ....................
V 2352 hCoV-19/Japan/PG-1823/2020|EPI_ISL_685429|2020-04 .................... .................... .................... .................... .................... ....................
V 2353 hCoV-19/Japan/PG-1824/2020|EPI_ISL_685430|2020-04 .................... .................... .................... .................... .................... ....................
V 2354 hCoV-19/Japan/PG-1825/2020|EPI_ISL_685431|2020-04 .................... .................... .................... .................... .................... ....................
V 2355 hCoV-19/Japan/PG-1826/2020|EPI_ISL_685432|2020-04 .................... .................... .................... .................... .................... ....................
V 2356 hCoV-19/Japan/PG-1827/2020|EPI_ISL_685433|2020-04 .................... .................... .................... .................... .................... ....................
V 2357 hCoV-19/Japan/PG-1828/2020|EPI_ISL_685434|2020-04 .................... .................... .................... .................... .................... ....................
V 2358 hCoV-19/Japan/PG-1829/2020|EPI_ISL_685435|2020-04 .................... .................... .................... .................... .................... ....................
V 2359 hCoV-19/Japan/PG-1830/2020|EPI_ISL_685436|2020-04 .................... .................... .................... .................... .................... ....................
V 2360 hCoV-19/Japan/PG-1831/2020|EPI_ISL_685437|2020-04 .................... .................... .................... .................... .................... ....................
V 2361 hCoV-19/Japan/PG-1832/2020|EPI_ISL_685438|2020-04 .................... .................... .................... .................... .................... ....................
V 2362 hCoV-19/Japan/PG-1833/2020|EPI_ISL_685439|2020-04 .................... .................... .................... .................... .................... ....................
V 2363 hCoV-19/Japan/PG-7630/2020|EPI_ISL_686732|2020-08 .................... .................... .................... .................... .................... ....................
V 2364 hCoV-19/Japan/PG-7631/2020|EPI_ISL_686733|2020-08 .................... .................... .................... .................... .................... ....................
V 2365 hCoV-19/Japan/PG-7632/2020|EPI_ISL_686734|2020-08 .................... .................... .................... .................... .................... ....................
V 2366 hCoV-19/Japan/PG-7633/2020|EPI_ISL_686735|2020-08 .................... .................... .................... .................... .................... ....................
V 2367 hCoV-19/Japan/PG-7634/2020|EPI_ISL_686736|2020-08 .................... .................... .................... .................... .................... ....................
V 2368 hCoV-19/Japan/PG-7635/2020|EPI_ISL_686737|2020-08 .................... .................... .................... .................... .................... ....................
V 2369 hCoV-19/Japan/PG-7636/2020|EPI_ISL_686738|2020-08 .................... .................... .................... .................... .................... ....................
V 2370 hCoV-19/Japan/PG-7637/2020|EPI_ISL_686739|2020-08 .................... .................... .................... .................... .................... ....................
V 2371 hCoV-19/Japan/PG-7638/2020|EPI_ISL_686740|2020-08 .................... .................... .................... .................... .................... ....................
V 2372 hCoV-19/Japan/PG-7639/2020|EPI_ISL_686741|2020-08 .................... .................... .................... .................... .................... ....................
V 2373 hCoV-19/Japan/PG-7640/2020|EPI_ISL_686742|2020-08 .................... .................... .................... .................... .................... ....................
V 2374 hCoV-19/Japan/PG-7641/2020|EPI_ISL_686743|2020-08 .................... .................... .................... .................... .................... ....................
V 2375 hCoV-19/Japan/PG-7642/2020|EPI_ISL_686744|2020-08 .................... .................... .................... .................... .................... ....................

| | | | | | | | | |
|---|---|---|---|---|---|---|---|---|
| V | 2376 | hCoV-19/Japan/PG-7643/2020\|EPI_ISL_686745\|2020-08 | .................... | .................... | ..................... | .................... | .................... | .................... |
| V | 2377 | hCoV-19/Japan/PG-7644/2020\|EPI_ISL_686746\|2020-08 | .................... | .................... | ..................... | .................... | .................... | .................... |
| V | 2378 | hCoV-19/Japan/PG-7645/2020\|EPI_ISL_686747\|2020-08 | .................... | .................... | ..................... | .................... | .................... | .................... |
| V | 2379 | hCoV-19/Japan/PG-7646/2020\|EPI_ISL_686748\|2020-08 | .................... | .................... | ..................... | .................... | .................... | .................... |
| V | 2380 | hCoV-19/Japan/PG-7647/2020\|EPI_ISL_686749\|2020-08 | .................... | .................... | ..................... | .................... | .................... | .................... |
| V | 2381 | hCoV-19/Japan/PG-7648/2020\|EPI_ISL_686750\|2020-08 | .................... | .................... | ..................... | .................... | .................... | .................... |
| V | 2382 | hCoV-19/Japan/PG-7649/2020\|EPI_ISL_686751\|2020-08 | .................... | .................... | ..................... | .................... | .................... | .................... |
| V | 2383 | hCoV-19/Japan/PG-7650/2020\|EPI_ISL_686752\|2020-08 | .................... | .................... | ..................... | .................... | .................... | .................... |
| V | 2384 | hCoV-19/Japan/PG-7651/2020\|EPI_ISL_686753\|2020-08 | .................... | .................... | ..................... | .................... | .................... | .................... |
| V | 2385 | hCoV-19/Japan/PG-7652/2020\|EPI_ISL_686754\|2020-08 | .................... | .................... | ..................... | .................... | .................... | .................... |
| V | 2386 | hCoV-19/Japan/PG-7653/2020\|EPI_ISL_686755\|2020-08 | .................... | .................... | ..................... | .................... | .................... | .................... |
| V | 2387 | hCoV-19/Japan/PG-7654/2020\|EPI_ISL_686756\|2020-08 | .................... | .................... | ..................... | .................... | .................... | .................... |
| V | 2388 | hCoV-19/Japan/PG-7655/2020\|EPI_ISL_686757\|2020-08 | .................... | .................... | ..................... | .................... | .................... | .................... |
| V | 2389 | hCoV-19/Japan/PG-7656/2020\|EPI_ISL_686758\|2020-08 | .................... | .................... | ..................... | .................... | .................... | .................... |
| V | 2390 | hCoV-19/Japan/PG-7657/2020\|EPI_ISL_686759\|2020-08 | .................... | .................... | ..................... | .................... | .................... | .................... |
| V | 2391 | hCoV-19/Japan/PG-7658/2020\|EPI_ISL_686760\|2020-08 | .................... | .................... | ..................... | .................... | .................... | .................... |
| V | 2392 | hCoV-19/Japan/PG-7659/2020\|EPI_ISL_686761\|2020-08 | .................... | .................... | ..................... | .................... | .................... | .................... |
| V | 2393 | hCoV-19/Japan/PG-7660/2020\|EPI_ISL_686762\|2020-08 | .................... | .................... | ..................... | .................... | .................... | .................... |
| V | 2394 | hCoV-19/Japan/PG-7661/2020\|EPI_ISL_686763\|2020-08 | .................... | .................... | ..................... | .................... | .................... | .................... |
| V | 2395 | hCoV-19/Japan/PG-7662/2020\|EPI_ISL_686764\|2020-08 | .................... | .A.................. | ..................... | .................... | .................... | .................... |
| V | 2396 | hCoV-19/Japan/PG-7663/2020\|EPI_ISL_686765\|2020-08 | .................... | .................... | ..................... | .................... | .................... | .................... |
| V | 2397 | hCoV-19/Japan/PG-7664/2020\|EPI_ISL_686766\|2020-08 | .................... | .................... | ..................... | .................... | .................... | .................... |
| V | 2398 | hCoV-19/Japan/PG-7665/2020\|EPI_ISL_686767\|2020-08 | .................... | .................... | ..................... | .................... | .................... | .................... |
| V | 2399 | hCoV-19/Japan/PG-7666/2020\|EPI_ISL_686768\|2020-08 | .................... | .................... | ..................... | .................... | .................... | .................... |
| V | 2400 | hCoV-19/Japan/PG-7667/2020\|EPI_ISL_686769\|2020-08 | .................... | .................... | ..................... | .................... | .................... | .................... |
| V | 2401 | hCoV-19/Japan/PG-3077/2020\|EPI_ISL_691200\|2020-04 | .................... | .................... | ..................... | .................... | .................... | .................... |
| V | 2402 | hCoV-19/Japan/PG-3078/2020\|EPI_ISL_691201\|2020-04 | .................... | .................... | ..................... | .................... | .................... | .................... |
| V | 2403 | hCoV-19/Japan/PG-3079/2020\|EPI_ISL_691202\|2020-04 | .................... | .................... | ..................... | .................... | .................... | .................... |
| V | 2404 | hCoV-19/Japan/PG-3080/2020\|EPI_ISL_691203\|2020-04 | .................... | .................... | ..................... | .................... | .................... | .................... |
| V | 2405 | hCoV-19/Japan/PG-3081/2020\|EPI_ISL_691204\|2020-04 | .................... | .................... | ..................... | .................... | .................... | .................... |
| V | 2406 | hCoV-19/Japan/PG-3082/2020\|EPI_ISL_691205\|2020-04 | .................... | .................... | ..................... | .................... | .................... | .................... |
| V | 2407 | hCoV-19/Japan/PG-3083/2020\|EPI_ISL_691206\|2020-04 | .................... | .................... | ..................... | .................... | .................... | .................... |
| V | 2408 | hCoV-19/Japan/PG-3084/2020\|EPI_ISL_691207\|2020-04 | .................... | .................... | ..................... | .................... | .................... | .................... |
| V | 2409 | hCoV-19/Japan/PG-3085/2020\|EPI_ISL_691208\|2020-04 | .................... | .................... | ..................... | .................... | .................... | .................... |
| V | 2410 | hCoV-19/Japan/PG-3086/2020\|EPI_ISL_691209\|2020-04 | .................... | .................... | ..................... | .................... | .................... | .................... |
| V | 2411 | hCoV-19/Japan/PG-3088/2020\|EPI_ISL_691210\|2020-04 | .................... | .................... | ..................... | .................... | .................... | .................... |
| V | 2412 | hCoV-19/Japan/PG-3089/2020\|EPI_ISL_691211\|2020-04 | .................... | .................... | ..................... | .................... | .................... | .................... |
| V | 2413 | hCoV-19/Japan/PG-3090/2020\|EPI_ISL_691212\|2020-04 | .................... | .................... | ..................... | .................... | .................... | .................... |
| V | 2414 | hCoV-19/Japan/PG-3091/2020\|EPI_ISL_691213\|2020-04 | .................... | .................... | ..................... | .................... | .................... | .................... |
| V | 2415 | hCoV-19/Japan/PG-3092/2020\|EPI_ISL_691214\|2020-04 | .................... | .................... | ..................... | .................... | .................... | .................... |
| V | 2416 | hCoV-19/Japan/PG-3093/2020\|EPI_ISL_691215\|2020-04 | .................... | .................... | ..................... | .................... | .................... | .................... |
| V | 2417 | hCoV-19/Japan/PG-3094/2020\|EPI_ISL_691216\|2020-04 | .................... | .................... | ..................... | .................... | .................... | .................... |
| V | 2418 | hCoV-19/Japan/PG-3095/2020\|EPI_ISL_691217\|2020-04 | .................... | .................... | ..................... | .................... | .................... | .................... |
| V | 2419 | hCoV-19/Japan/PG-3096/2020\|EPI_ISL_691218\|2020-04 | .................... | .................... | ..................... | .................... | .................... | .................... |
| V | 2420 | hCoV-19/Japan/PG-3097/2020\|EPI_ISL_691219\|2020-04 | .................... | .................... | ..................... | .................... | .................... | .................... |
| V | 2421 | hCoV-19/Japan/PG-3098/2020\|EPI_ISL_691220\|2020-04 | .................... | .................... | ..................... | .................... | .................... | .................... |
| V | 2422 | hCoV-19/Japan/PG-3099/2020\|EPI_ISL_691221\|2020-04 | .................... | .................... | ..................... | .................... | .................... | .................... |
| V | 2423 | hCoV-19/Japan/PG-3100/2020\|EPI_ISL_691222\|2020-04 | .................... | .................... | ..................... | .................... | .................... | .................... |
| V | 2424 | hCoV-19/Japan/PG-3101/2020\|EPI_ISL_691223\|2020-04 | .................... | .................... | ..................... | .................... | .................... | .................... |
| V | 2425 | hCoV-19/Japan/PG-3104/2020\|EPI_ISL_691226\|2020-05 | .................... | .................... | ..................... | .................... | .................... | .................... |
| V | 2426 | hCoV-19/Japan/PG-3106/2020\|EPI_ISL_691228\|2020-04 | .................... | .................... | ..................... | .................... | .................... | .................... |
| V | 2427 | hCoV-19/Japan/PG-3107/2020\|EPI_ISL_691229\|2020-04 | .................... | .................... | ..................... | .................... | .................... | .................... |
| V | 2428 | hCoV-19/Japan/PG-3170/2020\|EPI_ISL_691290\|2020-03 | .................... | .................... | ..................... | .................... | .................... | .................... |
| V | 2429 | hCoV-19/Japan/PG-3172/2020\|EPI_ISL_691292\|2020-03 | .................... | .................... | ..................... | .................... | .................... | .................... |
| V | 2430 | hCoV-19/Japan/PG-3173/2020\|EPI_ISL_691293\|2020-03 | .................... | .................... | ..................... | .................... | .................... | .................... |
| V | 2431 | hCoV-19/Japan/PG-3174/2020\|EPI_ISL_691294\|2020-03 | .................... | .................... | ..................... | .................... | .................... | .................... |
| V | 2432 | hCoV-19/Japan/PG-3175/2020\|EPI_ISL_691295\|2020-03 | .................... | .................... | ..................... | .................... | .................... | .................... |
| V | 2433 | hCoV-19/Japan/PG-3176/2020\|EPI_ISL_691296\|2020-03 | .................... | .................... | ..................... | .................... | .................... | .................... |
| V | 2434 | hCoV-19/Japan/PG-3177/2020\|EPI_ISL_691297\|2020-03 | .................... | .................... | ..................... | .................... | .................... | .................... |
| V | 2435 | hCoV-19/Japan/PG-3178/2020\|EPI_ISL_691298\|2020-03 | .................... | .................... | ..................... | .................... | .................... | .................... |
| V | 2436 | hCoV-19/Japan/PG-3179/2020\|EPI_ISL_691299\|2020-03 | .................... | .................... | ..................... | .................... | .................... | .................... |
| V | 2437 | hCoV-19/Japan/PG-4270/2020\|EPI_ISL_692500\|2020-07 | .................... | .................... | ..................... | .................... | .................... | .................... |
| V | 2438 | hCoV-19/Japan/PG-4271/2020\|EPI_ISL_692501\|2020-07 | .................... | .................... | ..................... | .................... | .................... | .................... |
| V | 2439 | hCoV-19/Japan/PG-4272/2020\|EPI_ISL_692502\|2020-07 | .................... | .................... | ..................... | .................... | .................... | .................... |
| V | 2440 | hCoV-19/Japan/PG-4273/2020\|EPI_ISL_692503\|2020-07 | .................... | .................... | ..................... | .................... | .................... | .................... |
| V | 2441 | hCoV-19/Japan/PG-4274/2020\|EPI_ISL_692504\|2020-07 | .................... | .................... | ..................... | .................... | .................... | .................... |
| V | 2442 | hCoV-19/Japan/PG-4275/2020\|EPI_ISL_692505\|2020-07 | .................... | .................... | ..................... | .................... | .................... | .................... |
| V | 2443 | hCoV-19/Japan/PG-4276/2020\|EPI_ISL_692506\|2020-07 | .................... | .................... | ..................... | .................... | .................... | .................... |
| V | 2444 | hCoV-19/Japan/PG-4277/2020\|EPI_ISL_692507\|2020-07 | .................... | .................... | ..................... | .................... | .................... | .................... |
| V | 2445 | hCoV-19/Japan/PG-4278/2020\|EPI_ISL_692508\|2020-07 | .................... | .................... | ..................... | .................... | .................... | .................... |
| V | 2446 | hCoV-19/Japan/PG-4279/2020\|EPI_ISL_692509\|2020-07 | .................... | .................... | ..................... | .................... | .................... | .................... |
| V | 2447 | hCoV-19/Japan/PG-4281/2020\|EPI_ISL_692510\|2020-07 | .................... | .................... | ..................... | .................... | .................... | .................... |
| V | 2448 | hCoV-19/Japan/PG-4282/2020\|EPI_ISL_692511\|2020-07 | .................... | .................... | ..................... | .................... | .................... | .................... |
| V | 2449 | hCoV-19/Japan/PG-4283/2020\|EPI_ISL_692512\|2020-07 | .................... | .................... | ..................... | .................... | .................... | .................... |
| V | 2450 | hCoV-19/Japan/PG-4284/2020\|EPI_ISL_692513\|2020-07 | .................... | .................... | ..................... | .................... | .................... | .................... |
| V | 2451 | hCoV-19/Japan/PG-4285/2020\|EPI_ISL_692514\|2020-07 | .................... | .................... | ..................... | .................... | .................... | .................... |
| V | 2452 | hCoV-19/Japan/PG-4286/2020\|EPI_ISL_692515\|2020-07 | .................... | .................... | ..................... | .................... | .................... | .................... |
| V | 2453 | hCoV-19/Japan/PG-4287/2020\|EPI_ISL_692516\|2020-07 | .................... | .................... | ..................... | .................... | .................... | .................... |
| V | 2454 | hCoV-19/Japan/PG-4288/2020\|EPI_ISL_692517\|2020-07 | .................... | .................... | ..................... | .................... | .................... | .................... |
| V | 2455 | hCoV-19/Japan/PG-4289/2020\|EPI_ISL_692518\|2020-07 | .................... | .................... | ..................... | .................... | .................... | .................... |

| | | | | | | | |
|---|---|---|---|---|---|---|---|
| V | 2456 hCoV-19/Japan/PG-4290/2020\|EPI_ISL_692519\|2020-07 | ···················· | ···················· | ····················· | ···················· | ···················· | ···················· |
| V | 2457 hCoV-19/Japan/PG-4291/2020\|EPI_ISL_692520\|2020-07 | ···················· | ···················· | ····················· | ···················· | ···················· | ···················· |
| V | 2458 hCoV-19/Japan/PG-4292/2020\|EPI_ISL_692521\|2020-07 | ···················· | ···················· | ····················· | ···················· | ···················· | ···················· |
| V | 2459 hCoV-19/Japan/PG-4293/2020\|EPI_ISL_692522\|2020-07 | ···················· | ···················· | ····················· | ···················· | ···················· | ···················· |
| V | 2460 hCoV-19/Japan/PG-4294/2020\|EPI_ISL_692523\|2020-07 | ···················· | ···················· | ····················· | ···················· | ···················· | ···················· |
| V | 2461 hCoV-19/Japan/PG-4295/2020\|EPI_ISL_692524\|2020-07 | ···················· | ···················· | ····················· | ···················· | ···················· | ···················· |
| V | 2462 hCoV-19/Japan/PG-4296/2020\|EPI_ISL_692525\|2020-07 | ···················· | ···················· | ····················· | ···················· | ···················· | ···················· |
| V | 2463 hCoV-19/Japan/PG-4297/2020\|EPI_ISL_692526\|2020-07 | ···················· | ···················· | ····················· | ···················· | ···················· | ···················· |
| V | 2464 hCoV-19/Japan/PG-4298/2020\|EPI_ISL_692527\|2020-05 | ···················· | ···················· | ····················· | ···················· | ···················· | ···················· |
| V | 2465 hCoV-19/Japan/PG-4299/2020\|EPI_ISL_692528\|2020-06 | ···················· | ···················· | ····················· | ···················· | ···················· | ···················· |
| V | 2466 hCoV-19/Japan/PG-4301/2020\|EPI_ISL_692529\|2020-07 | ···················· | ···················· | ····················· | ···················· | ···················· | ···················· |
| V | 2467 hCoV-19/Japan/PG-4302/2020\|EPI_ISL_692530\|2020-07 | ···················· | ···················· | ····················· | ···················· | ···················· | ···················· |
| V | 2468 hCoV-19/Japan/PG-4303/2020\|EPI_ISL_692531\|2020-07 | ···················· | ···················· | ····················· | ···················· | ···················· | ···················· |
| V | 2469 hCoV-19/Japan/PG-4304/2020\|EPI_ISL_692532\|2020-07 | ···················· | ···················· | ····················· | ···················· | ···················· | ···················· |
| V | 2470 hCoV-19/Japan/PG-4305/2020\|EPI_ISL_692533\|2020-07 | ···················· | ···················· | ····················· | ···················· | ···················· | ···················· |
| V | 2471 hCoV-19/Japan/PG-4306/2020\|EPI_ISL_692534\|2020-07 | ···················· | ···················· | ····················· | ···················· | ···················· | ···················· |
| V | 2472 hCoV-19/Japan/PG-4307/2020\|EPI_ISL_692535\|2020-07 | ···················· | ···················· | ····················· | ···················· | ···················· | ···················· |
| V | 2473 hCoV-19/Japan/PG-4308/2020\|EPI_ISL_692536\|2020-07 | ···················· | ···················· | ····················· | ···················· | ···················· | ···················· |
| V | 2474 hCoV-19/Japan/PG-4309/2020\|EPI_ISL_692537\|2020-07 | ···················· | ···················· | ····················· | ···················· | ···················· | ···················· |
| V | 2475 hCoV-19/Japan/PG-4310/2020\|EPI_ISL_692538\|2020-07 | ···················· | ···················· | ····················· | ···················· | ···················· | ···················· |
| V | 2476 hCoV-19/Japan/PG-4311/2020\|EPI_ISL_692539\|2020-07 | ···················· | ···················· | ····················· | ···················· | ···················· | ···················· |
| V | 2477 hCoV-19/Japan/PG-4312/2020\|EPI_ISL_692540\|2020-07 | ···················· | ···················· | ····················· | ···················· | ···················· | ···················· |
| V | 2478 hCoV-19/Japan/PG-4313/2020\|EPI_ISL_692541\|2020-07 | ···················· | ···················· | ····················· | ···················· | ···················· | ···················· |
| V | 2479 hCoV-19/Japan/PG-4314/2020\|EPI_ISL_692542\|2020-07 | ···················· | ···················· | ····················· | ···················· | ···················· | ···················· |
| V | 2480 hCoV-19/Japan/PG-4315/2020\|EPI_ISL_692543\|2020-07 | ···················· | ···················· | ····················· | ···················· | ···················· | ···················· |
| V | 2481 hCoV-19/Japan/PG-4316/2020\|EPI_ISL_692544\|2020-07 | ···················· | ···················· | ····················· | ···················· | ···················· | ···················· |
| V | 2482 hCoV-19/Japan/PG-4317/2020\|EPI_ISL_692545\|2020-07 | ···················· | ···················· | ····················· | ···················· | ···················· | ···················· |
| V | 2483 hCoV-19/Japan/PG-4318/2020\|EPI_ISL_692546\|2020-07 | ···················· | ···················· | ····················· | ···················· | ···················· | ···················· |
| V | 2484 hCoV-19/Japan/PG-4319/2020\|EPI_ISL_692547\|2020-07 | ···················· | ···················· | ····················· | ···················· | ···················· | ···················· |
| V | 2485 hCoV-19/Japan/PG-4320/2020\|EPI_ISL_692548\|2020-07 | ···················· | ···················· | ····················· | ···················· | ···················· | ···················· |
| V | 2486 hCoV-19/Japan/PG-4321/2020\|EPI_ISL_692549\|2020-07 | ···················· | ···················· | ····················· | ···················· | ···················· | ···················· |
| V | 2487 hCoV-19/Japan/PG-4322/2020\|EPI_ISL_692550\|2020-07 | ···················· | ···················· | ····················· | ···················· | ···················· | ···················· |
| V | 2488 hCoV-19/Japan/PG-4323/2020\|EPI_ISL_692551\|2020-07 | ···················· | ···················· | ····················· | ···················· | ···················· | ···················· |
| V | 2489 hCoV-19/Japan/PG-4324/2020\|EPI_ISL_692552\|2020-07 | ···················· | ···················· | ····················· | ···················· | ···················· | ···················· |
| V | 2490 hCoV-19/Japan/PG-4325/2020\|EPI_ISL_692553\|2020-07 | ···················· | ···················· | ····················· | ···················· | ···················· | ···················· |
| V | 2491 hCoV-19/Japan/PG-4326/2020\|EPI_ISL_692554\|2020-07 | ···················· | ···················· | ····················· | ···················· | ···················· | ···················· |
| V | 2492 hCoV-19/Japan/PG-4329/2020\|EPI_ISL_692557\|2020-07 | ···················· | ···················· | ····················· | ···················· | ···················· | ···················· |
| V | 2493 hCoV-19/Japan/PG-4330/2020\|EPI_ISL_692558\|2020-07 | ···················· | ···················· | ····················· | ···················· | ···················· | ···················· |
| V | 2494 hCoV-19/Japan/PG-4331/2020\|EPI_ISL_692559\|2020-07 | ···················· | ···················· | ····················· | ···················· | ···················· | ···················· |
| V | 2495 hCoV-19/Switzerland/TI-ETHZ-350121/2020\|EPI_ISL_69 | ···················· | ···················· | ····················· | ···················· | ···················· | ···················· |
| V | 2496 hCoV-19/Switzerland/LU-ETHZ-350123/2020\|EPI_ISL_69 | ···················· | ···················· | ····················· | ···················· | ···················· | ···················· |
| V | 2497 hCoV-19/Switzerland/ZH-ETHZ-350124/2020\|EPI_ISL_69 | ···················· | ···················· | ····················· | ···················· | ···················· | ···················· |
| V | 2498 hCoV-19/Switzerland/AG-ETHZ-350125/2020\|EPI_ISL_69 | ···················· | ···················· | ····················· | ···················· | ···················· | ···················· |
| V | 2499 hCoV-19/Switzerland/ZH-ETHZ-350126/2020\|EPI_ISL_69 | ···················· | ·A·················· | ····················· | ···················· | ···················· | ···················· |
| V | 2500 hCoV-19/Switzerland/AG-ETHZ-350127/2020\|EPI_ISL_69 | ···················· | ···················· | ····················· | ···················· | ···················· | ···················· |
| V | 2501 hCoV-19/Switzerland/BE-ETHZ-350175/2020\|EPI_ISL_69 | ···················· | ···················· | ····················· | ···················· | ···················· | ···················· |
| V | 2502 hCoV-19/Switzerland/SH-ETHZ-350176/2020\|EPI_ISL_69 | ···················· | ···················· | ····················· | ···················· | ·····K·············· | ···················· |
| V | 2503 hCoV-19/Switzerland/ZH-ETHZ-350177/2020\|EPI_ISL_69 | ···················· | ···················· | ····················· | ···················· | ···················· | ···················· |
| V | 2504 hCoV-19/Switzerland/ZH-ETHZ-350178/2020\|EPI_ISL_69 | ···················· | ···················· | ····················· | ···················· | ···················· | ···················· |
| V | 2505 hCoV-19/Switzerland/BE-ETHZ-350179/2020\|EPI_ISL_69 | ···················· | ···················· | ····················· | ···················· | ···················· | ···················· |
| V | 2506 hCoV-19/Switzerland/SO-ETHZ-350181/2020\|EPI_ISL_69 | ···················· | ···················· | ····················· | ···················· | ···················· | ···················· |
| V | 2507 hCoV-19/Switzerland/BE-ETHZ-350183/2020\|EPI_ISL_69 | ···················· | ···················· | ····················· | ···················· | ···················· | ···················· |
| V | 2508 hCoV-19/Switzerland/SH-ETHZ-350184/2020\|EPI_ISL_69 | ···T················ | ···················· | ····················· | ···················· | ·····K·············· | ···················· |
| V | 2509 hCoV-19/Switzerland/SO-ETHZ-350185/2020\|EPI_ISL_69 | ···················· | ···················· | ····················· | ···················· | ···················· | ···················· |
| V | 2510 hCoV-19/Switzerland/AG-ETHZ-350186/2020\|EPI_ISL_69 | ···················· | ···················· | ····················· | ···················· | ···················· | ···················· |
| V | 2511 hCoV-19/Switzerland/VD-ETHZ-350187/2020\|EPI_ISL_69 | ···················· | ···················· | ····················· | ···················· | ···················· | ···················· |
| V | 2512 hCoV-19/Switzerland/SH-ETHZ-350188/2020\|EPI_ISL_69 | ···················· | ···················· | ····················· | ···················· | ···················· | ···················· |
| V | 2513 hCoV-19/Switzerland/ZH-ETHZ-350189/2020\|EPI_ISL_69 | ···················· | ···················· | ····················· | ···················· | ···················· | ···················· |
| V | 2514 hCoV-19/Switzerland/BE-ETHZ-350190/2020\|EPI_ISL_69 | ···················· | ···················· | ····················· | ···················· | ···················· | ···················· |
| V | 2515 hCoV-19/Switzerland/BE-ETHZ-350191/2020\|EPI_ISL_69 | ···················· | ···················· | ····················· | ···················· | ···················· | ···················· |
| V | 2516 hCoV-19/Switzerland/VD-ETHZ-350192/2020\|EPI_ISL_69 | ···················· | ···················· | ····················· | ···················· | ···················· | ···················· |
| V | 2517 hCoV-19/Switzerland/BE-ETHZ-350193/2020\|EPI_ISL_69 | ···················· | ···················· | ····················· | ···················· | ···················· | ···················· |
| V | 2518 hCoV-19/Switzerland/NE-ETHZ-350194/2020\|EPI_ISL_69 | ···················· | ···················· | ····················· | ···················· | ···················· | ···················· |
| V | 2519 hCoV-19/Switzerland/VD-ETHZ-350195/2020\|EPI_ISL_69 | ···················· | ···················· | ····················· | ···················· | ···················· | ···················· |
| V | 2520 hCoV-19/Switzerland/VD-ETHZ-350197/2020\|EPI_ISL_69 | ···················· | ···················· | ····················· | ···················· | ···················· | ···················· |
| V | 2521 hCoV-19/Switzerland/BE-ETHZ-350198/2020\|EPI_ISL_69 | ···················· | ···················· | ····················· | ···················· | ···················· | ···················· |
| V | 2522 hCoV-19/Switzerland/VD-ETHZ-350199/2020\|EPI_ISL_69 | ···················· | ···················· | ····················· | ···················· | ···················· | ···················· |
| V | 2523 hCoV-19/Switzerland/ZH-ETHZ-350200/2020\|EPI_ISL_69 | ···················· | ···················· | ····················· | ···················· | ···················· | ···················· |
| V | 2524 hCoV-19/Switzerland/GE-ETHZ-350201/2020\|EPI_ISL_69 | ···················· | ···················· | ····················· | ···················· | ···················· | ···················· |
| V | 2525 hCoV-19/Switzerland/VD-ETHZ-350202/2020\|EPI_ISL_69 | ···················· | ···················· | ····················· | ···················· | ···················· | ···················· |
| V | 2526 hCoV-19/Switzerland/BE-ETHZ-350203/2020\|EPI_ISL_69 | ···················· | ···················· | ····················· | ···················· | ···················· | ···················· |
| V | 2527 hCoV-19/Switzerland/ZH-ETHZ-350204/2020\|EPI_ISL_69 | ···················· | ···················· | ····················· | ···················· | ···················· | ···················· |
| V | 2528 hCoV-19/Switzerland/VD-ETHZ-350205/2020\|EPI_ISL_69 | ···················· | ···················· | ····················· | ···················· | ···················· | ·R·················· |
| V | 2529 hCoV-19/Switzerland/GE-ETHZ-350206/2020\|EPI_ISL_69 | ···················· | ···················· | ····················· | ···················· | ···················· | ···················· |
| V | 2530 hCoV-19/Switzerland/GE-ETHZ-350207/2020\|EPI_ISL_69 | ···················· | ···M················ | ····················· | ···················· | ···················· | ···················· |
| V | 2531 hCoV-19/Switzerland/GE-ETHZ-350208/2020\|EPI_ISL_69 | ···················· | ···················· | ····················· | ···················· | ···················· | ···················· |
| V | 2532 hCoV-19/Switzerland/VD-ETHZ-350209/2020\|EPI_ISL_69 | ···················· | ···················· | ····················· | ···················· | ···················· | ···················· |
| V | 2533 hCoV-19/Switzerland/GE-ETHZ-350211/2020\|EPI_ISL_69 | ···················· | ···················· | ····················· | ···················· | ···················· | ···················· |
| V | 2534 hCoV-19/Switzerland/AG-ETHZ-360000/2020\|EPI_ISL_69 | ···················· | ···················· | ····················· | ···················· | ·············Y······ | ···················· |
| V | 2535 hCoV-19/Switzerland/BL-ETHZ-360001/2020\|EPI_ISL_69 | ···················· | ···················· | ····················· | ···················· | ···················· | ···················· |

| | | | | | | | | |
|---|---|---|---|---|---|---|---|---|
| V | 2536 | hCoV-19/Switzerland/BS-ETHZ-360002/2020\|EPI_ISL_69 | ···················· | ···················· | ···················· | ···················· | ···················· | ···················· |
| V | 2537 | hCoV-19/Switzerland/BL-ETHZ-360003/2020\|EPI_ISL_69 | ···················· | ···················· | ···················· | ···················· | ···················· | ···················· |
| V | 2538 | hCoV-19/Switzerland/BL-ETHZ-360004/2020\|EPI_ISL_69 | ···················· | ···················· | ···················· | ···················· | ···················· | ···················· |
| V | 2539 | hCoV-19/Switzerland/BL-ETHZ-360005/2020\|EPI_ISL_69 | ···················· | ·A·················· | ···················· | ···················· | ···················· | ··A················· |
| V | 2540 | hCoV-19/Switzerland/BL-ETHZ-360006/2020\|EPI_ISL_69 | ···················· | ·A·················· | ···················· | ···················· | ···················· | ···················· |
| V | 2541 | hCoV-19/Switzerland/BS-ETHZ-360007/2020\|EPI_ISL_69 | ···················· | ···················· | ···················· | ···················· | ···················· | ···················· |
| V | 2542 | hCoV-19/Switzerland/BL-ETHZ-360008/2020\|EPI_ISL_69 | ···················· | ···················· | ···················· | ···················· | ···················· | ···················· |
| V | 2543 | hCoV-19/Switzerland/BL-ETHZ-360009/2020\|EPI_ISL_69 | ···················· | ·A·················· | ···················· | ···················· | ···················· | ···················· |
| V | 2544 | hCoV-19/Switzerland/BS-ETHZ-360010/2020\|EPI_ISL_69 | ···················· | ···················· | ···················· | ···················· | ···················· | ···················· |
| V | 2545 | hCoV-19/Switzerland/BL-ETHZ-360012/2020\|EPI_ISL_69 | ···················· | ···················· | ···················· | ···················· | ···················· | ···················· |
| V | 2546 | hCoV-19/Switzerland/BS-ETHZ-360013/2020\|EPI_ISL_69 | ···················· | ···················· | ···················· | ···················· | ···················· | ···················· |
| V | 2547 | hCoV-19/Switzerland/BS-ETHZ-360014/2020\|EPI_ISL_69 | ···················· | ···················· | ···················· | ···················· | ···················· | ···················· |
| V | 2548 | hCoV-19/Switzerland/SO-ETHZ-360015/2020\|EPI_ISL_69 | ···················· | ·A·················· | ···················· | ···················· | ···················· | ···················· |
| V | 2549 | hCoV-19/Switzerland/BE-ETHZ-360017/2020\|EPI_ISL_69 | ···················· | ···················· | ···················· | ···················· | ···················· | ··R················· |
| V | 2550 | hCoV-19/Switzerland/BE-ETHZ-360018/2020\|EPI_ISL_69 | ···················· | ···················· | ···················· | ···················· | ···················· | ···················· |
| V | 2551 | hCoV-19/Switzerland/BE-ETHZ-360019/2020\|EPI_ISL_69 | ···················· | ·········A·········· | ···················· | ···················· | ···················· | ···················· |
| V | 2552 | hCoV-19/Switzerland/BE-ETHZ-360022/2020\|EPI_ISL_69 | ···················· | ···················· | ···················· | ···················· | ···················· | ···················· |
| V | 2553 | hCoV-19/Switzerland/BE-ETHZ-360023/2020\|EPI_ISL_69 | ···················· | ·A·················· | ···················· | ···················· | ···················· | ···················· |
| V | 2554 | hCoV-19/Switzerland/BE-ETHZ-360024/2020\|EPI_ISL_69 | ···················· | ···················· | ···················· | ···················· | ···················· | ···················· |
| V | 2555 | hCoV-19/Switzerland/BE-ETHZ-360025/2020\|EPI_ISL_69 | ···················· | ···················· | ···················· | ···················· | ···················· | ···················· |
| V | 2556 | hCoV-19/Switzerland/BE-ETHZ-360028/2020\|EPI_ISL_69 | ···················· | ···················· | ···················· | ···················· | ···················· | ···················· |
| V | 2557 | hCoV-19/Switzerland/BE-ETHZ-360031/2020\|EPI_ISL_69 | ···················· | ···················· | ···················· | ···················· | ···················· | ···················· |
| V | 2558 | hCoV-19/Switzerland/BE-ETHZ-360032/2020\|EPI_ISL_69 | ···················· | ···················· | ···················· | ···················· | ···················· | ···················· |
| V | 2559 | hCoV-19/Switzerland/VD-ETHZ-360047/2020\|EPI_ISL_69 | ···················· | ···················· | ···················· | ···················· | ···················· | ···················· |
| V | 2560 | hCoV-19/Switzerland/GR-ETHZ-360065/2020\|EPI_ISL_69 | ···················· | ···················· | ···················· | ···················· | ···················· | ···················· |
| V | 2561 | hCoV-19/South Africa/UFS-VIRO-NGS-38/2020\|EPI_ISL_ | ···················· | ·A·················· | ···················· | ···················· | ···················· | ···················· |
| V | 2562 | hCoV-19/Japan/PG-1220/2020\|EPI_ISL_684830\|2020-04 | ···················· | ···················· | ···················· | ···················· | ···················· | ···················· |
| V | 2563 | hCoV-19/Japan/PG-1221/2020\|EPI_ISL_684831\|2020-04 | ···················· | ···················· | ···················· | ···················· | ···················· | ···················· |
| V | 2564 | hCoV-19/Japan/PG-1222/2020\|EPI_ISL_684832\|2020-04 | ···················· | ···················· | ···················· | ···················· | ···················· | ···················· |
| V | 2565 | hCoV-19/Japan/PG-1223/2020\|EPI_ISL_684833\|2020-04 | ···················· | ···················· | ···················· | ···················· | ···················· | ···················· |
| V | 2566 | hCoV-19/Japan/PG-1224/2020\|EPI_ISL_684834\|2020-04 | ···················· | ···················· | ···················· | ···················· | ···················· | ···················· |
| V | 2567 | hCoV-19/Japan/PG-1225/2020\|EPI_ISL_684835\|2020-04 | ···················· | ···················· | ···················· | ···················· | ···················· | ···················· |
| V | 2568 | hCoV-19/Japan/PG-1226/2020\|EPI_ISL_684836\|2020-04 | ···················· | ···················· | ···················· | ···················· | ···················· | ···················· |
| V | 2569 | hCoV-19/Japan/PG-1227/2020\|EPI_ISL_684837\|2020-04 | ···················· | ···················· | ···················· | ···················· | ···················· | ···················· |
| V | 2570 | hCoV-19/Japan/PG-1228/2020\|EPI_ISL_684838\|2020-04 | ···················· | ···················· | ···················· | ···················· | ···················· | ···················· |
| V | 2571 | hCoV-19/Japan/PG-1229/2020\|EPI_ISL_684839\|2020-04 | ···················· | ···················· | ···················· | ···················· | ···················· | ···················· |
| V | 2572 | hCoV-19/Japan/PG-1230/2020\|EPI_ISL_684840\|2020-04 | ···················· | ···················· | ···················· | ···················· | ···················· | ···················· |
| V | 2573 | hCoV-19/Japan/PG-1231/2020\|EPI_ISL_684841\|2020-04 | ···················· | ···················· | ···················· | ···················· | ···················· | ···················· |
| V | 2574 | hCoV-19/Japan/PG-1232/2020\|EPI_ISL_684842\|2020-04 | ···················· | ···················· | ···················· | ···················· | ···················· | ···················· |
| V | 2575 | hCoV-19/Japan/PG-1233/2020\|EPI_ISL_684843\|2020-04 | ···················· | ···················· | ···················· | ···················· | ···················· | ···················· |
| V | 2576 | hCoV-19/Japan/PG-1234/2020\|EPI_ISL_684844\|2020-04 | ···················· | ···················· | ···················· | ···················· | ···················· | ···················· |
| V | 2577 | hCoV-19/Japan/PG-1235/2020\|EPI_ISL_684845\|2020-04 | ···················· | ···················· | ···················· | ···················· | ···················· | ···················· |
| V | 2578 | hCoV-19/Japan/PG-1236/2020\|EPI_ISL_684846\|2020-04 | ···················· | ···················· | ···················· | ···················· | ···················· | ···················· |
| V | 2579 | hCoV-19/Japan/PG-1237/2020\|EPI_ISL_684847\|2020-04 | ···················· | ···················· | ···················· | ···················· | ···················· | ···················· |
| V | 2580 | hCoV-19/Japan/PG-1238/2020\|EPI_ISL_684848\|2020-04 | ···················· | ···················· | ···················· | ···················· | ···················· | ···················· |
| V | 2581 | hCoV-19/Japan/PG-1239/2020\|EPI_ISL_684849\|2020-04 | ···················· | ···················· | ···················· | ···················· | ···················· | ···················· |
| V | 2582 | hCoV-19/Japan/PG-1240/2020\|EPI_ISL_684850\|2020-04 | ···················· | ···················· | ···················· | ···················· | ···················· | ···················· |
| V | 2583 | hCoV-19/Japan/PG-1241/2020\|EPI_ISL_684851\|2020-04 | ···················· | ···················· | ···················· | ···················· | ···················· | ···················· |
| V | 2584 | hCoV-19/Japan/PG-1242/2020\|EPI_ISL_684852\|2020-04 | ···················· | ···················· | ···················· | ···················· | ···················· | ···················· |
| V | 2585 | hCoV-19/Japan/PG-1243/2020\|EPI_ISL_684853\|2020-04 | ···················· | ···················· | ···················· | ···················· | ···················· | ···················· |
| V | 2586 | hCoV-19/Japan/PG-1244/2020\|EPI_ISL_684854\|2020-04 | ···················· | ···················· | ···················· | ···················· | ···················· | ···················· |
| V | 2587 | hCoV-19/Japan/PG-1245/2020\|EPI_ISL_684855\|2020-04 | ···················· | ···················· | ···················· | ···················· | ···················· | ···················· |
| V | 2588 | hCoV-19/Japan/PG-1246/2020\|EPI_ISL_684856\|2020-04 | ···················· | ···················· | ···················· | ···················· | ···················· | ···················· |
| V | 2589 | hCoV-19/Japan/PG-1247/2020\|EPI_ISL_684857\|2020 | ···················· | ···················· | ···················· | ···················· | ···················· | ···················· |
| V | 2590 | hCoV-19/Japan/PG-1248/2020\|EPI_ISL_684858\|2020 | ···················· | ···················· | ···················· | ···················· | ···················· | ···················· |
| V | 2591 | hCoV-19/Japan/PG-1249/2020\|EPI_ISL_684859\|2020 | ···················· | ···················· | ···················· | ···················· | ···················· | ···················· |
| V | 2592 | hCoV-19/Japan/PG-1250/2020\|EPI_ISL_684860\|2020 | ···················· | ···················· | ···················· | ···················· | ···················· | ···················· |
| V | 2593 | hCoV-19/Japan/PG-1251/2020\|EPI_ISL_684861\|2020 | ···················· | ···················· | ···················· | ···················· | ···················· | ···················· |
| V | 2594 | hCoV-19/Japan/PG-1252/2020\|EPI_ISL_684862\|2020 | ···················· | ···················· | ···················· | ···················· | ···················· | ···················· |
| V | 2595 | hCoV-19/Japan/PG-1253/2020\|EPI_ISL_684863\|2020 | ···················· | ···················· | ···················· | ···················· | ···················· | ···················· |
| V | 2596 | hCoV-19/Japan/PG-1254/2020\|EPI_ISL_684864\|2020 | ···················· | ···················· | ···················· | ···················· | ···················· | ···················· |
| V | 2597 | hCoV-19/Japan/PG-1255/2020\|EPI_ISL_684865\|2020 | ···················· | ···················· | ···················· | ···················· | ···················· | ···················· |
| V | 2598 | hCoV-19/Japan/PG-1256/2020\|EPI_ISL_684866\|2020 | ···················· | ···················· | ···················· | ···················· | ···················· | ···················· |
| V | 2599 | hCoV-19/Japan/PG-1257/2020\|EPI_ISL_684867\|2020 | ···················· | ···················· | ···················· | ···················· | ···················· | ···················· |
| V | 2600 | hCoV-19/Japan/PG-1258/2020\|EPI_ISL_684868\|2020 | ···················· | ···················· | ···················· | ···················· | ···················· | ···················· |
| V | 2601 | hCoV-19/Japan/PG-1259/2020\|EPI_ISL_684869\|2020 | ···················· | ···················· | ···················· | ···················· | ···················· | ···················· |
| V | 2602 | hCoV-19/Japan/PG-1260/2020\|EPI_ISL_684870\|2020 | ···················· | ···················· | ···················· | ···················· | ···················· | ···················· |
| V | 2603 | hCoV-19/Japan/PG-1261/2020\|EPI_ISL_684871\|2020 | ···················· | ···················· | ···················· | ···················· | ···················· | ···················· |
| V | 2604 | hCoV-19/Japan/PG-1262/2020\|EPI_ISL_684872\|2020 | ···················· | ···················· | ···················· | ···················· | ···················· | ···················· |
| V | 2605 | hCoV-19/Japan/PG-1263/2020\|EPI_ISL_684873\|2020 | ···················· | ···················· | ···················· | ···················· | ···················· | ···················· |
| V | 2606 | hCoV-19/Japan/PG-1264/2020\|EPI_ISL_684874\|2020 | ···················· | ···················· | ···················· | ···················· | ···················· | ···················· |
| V | 2607 | hCoV-19/Japan/PG-1265/2020\|EPI_ISL_684875\|2020 | ···················· | ···················· | ···················· | ···················· | ···················· | ···················· |
| V | 2608 | hCoV-19/Japan/PG-1266/2020\|EPI_ISL_684876\|2020 | ···················· | ···················· | ···················· | ···················· | ···················· | ···················· |
| V | 2609 | hCoV-19/Japan/PG-1267/2020\|EPI_ISL_684877\|2020 | ···················· | ···················· | ···················· | ···················· | ···················· | ···················· |
| V | 2610 | hCoV-19/Japan/PG-1268/2020\|EPI_ISL_684878\|2020 | ···················· | ···················· | ···················· | ···················· | ···················· | ···················· |
| V | 2611 | hCoV-19/Japan/PG-1269/2020\|EPI_ISL_684879\|2020 | ···················· | ···················· | ···················· | ···················· | ···················· | ···················· |
| V | 2612 | hCoV-19/Japan/PG-1270/2020\|EPI_ISL_684880\|2020 | ···················· | ···················· | ···················· | ···················· | ···················· | ···················· |
| V | 2613 | hCoV-19/Japan/PG-1272/2020\|EPI_ISL_684881\|2020-03 | ···················· | ···················· | ···················· | ···················· | ···················· | ···················· |
| V | 2614 | hCoV-19/Japan/PG-1273/2020\|EPI_ISL_684882\|2020-03 | ···················· | ···················· | ···················· | ···················· | ···················· | ···················· |
| V | 2615 | hCoV-19/Japan/PG-1274/2020\|EPI_ISL_684883\|2020-03 | ···················· | ···················· | ···················· | ···················· | ···················· | ···················· |

| | | | | | | | | |
|---|---|---|---|---|---|---|---|---|
| V | 2616 | hCoV-19/Japan/PG-1275/2020\|EPI_ISL_684884\|2020-03 | .................... | .................... | ..................... | .................... | .................... | .................... |
| V | 2617 | hCoV-19/Japan/PG-1276/2020\|EPI_ISL_684885\|2020-03 | .................... | .................... | ..................... | .................... | .................... | .................... |
| V | 2618 | hCoV-19/Japan/PG-1277/2020\|EPI_ISL_684886\|2020-03 | .................... | .................... | ..................... | .................... | .................... | .................... |
| V | 2619 | hCoV-19/Japan/PG-1278/2020\|EPI_ISL_684887\|2020-03 | .................... | .................... | ..................... | .................... | .................... | .................... |
| V | 2620 | hCoV-19/Japan/PG-1279/2020\|EPI_ISL_684888\|2020-03 | .................... | .................... | ..................... | .................... | .................... | .................... |
| V | 2621 | hCoV-19/Japan/PG-1280/2020\|EPI_ISL_684889\|2020-03 | .................... | .................... | ..................... | .................... | .................... | .................... |
| V | 2622 | hCoV-19/Japan/PG-1281/2020\|EPI_ISL_684890\|2020-03 | .................... | .................... | ..................... | .................... | .................... | .................... |
| V | 2623 | hCoV-19/Japan/PG-1282/2020\|EPI_ISL_684891\|2020-03 | .................... | .................... | ..................... | .................... | .................... | .................... |
| V | 2624 | hCoV-19/Japan/PG-1283/2020\|EPI_ISL_684892\|2020-03 | .................... | .................... | ..................... | .................... | .................... | .................... |
| V | 2625 | hCoV-19/Japan/PG-1284/2020\|EPI_ISL_684893\|2020-03 | .................... | .................... | ..................... | .................... | .................... | .................... |
| V | 2626 | hCoV-19/Japan/PG-1285/2020\|EPI_ISL_684894\|2020-03 | .................... | .................... | ..................... | .................... | .................... | .................... |
| V | 2627 | hCoV-19/Japan/PG-1286/2020\|EPI_ISL_684895\|2020-03 | .................... | .................... | ..................... | .................... | .................... | .................... |
| V | 2628 | hCoV-19/Japan/PG-1287/2020\|EPI_ISL_684896\|2020-03 | .................... | .................... | ..................... | .................... | .................... | .................... |
| V | 2629 | hCoV-19/Japan/PG-1288/2020\|EPI_ISL_684897\|2020-03 | .................... | .................... | ..................... | .................... | .................... | .................... |
| V | 2630 | hCoV-19/Japan/PG-1290/2020\|EPI_ISL_684898\|2020-03 | .................... | .................... | ..................... | .................... | .................... | .................... |
| V | 2631 | hCoV-19/Japan/PG-1291/2020\|EPI_ISL_684899\|2020-03 | .................... | .................... | ..................... | .................... | .................... | .................... |
| V | 2632 | hCoV-19/Japan/PG-1762/2020\|EPI_ISL_685368\|2020-04 | .................... | .................... | ..................... | .................... | .................... | .................... |
| V | 2633 | hCoV-19/Japan/PG-1763/2020\|EPI_ISL_685369\|2020-04 | .................... | .................... | ..................... | .................... | .................... | .................... |
| V | 2634 | hCoV-19/Japan/PG-1764/2020\|EPI_ISL_685370\|2020-04 | .................... | .................... | ..................... | .................... | .................... | .................... |
| V | 2635 | hCoV-19/Japan/PG-1765/2020\|EPI_ISL_685371\|2020-04 | .................... | .................... | ..................... | .................... | .................... | .................... |
| V | 2636 | hCoV-19/Japan/PG-1766/2020\|EPI_ISL_685372\|2020-04 | .................... | .................... | ..................... | .................... | .................... | .................... |
| V | 2637 | hCoV-19/Japan/PG-1767/2020\|EPI_ISL_685373\|2020-04 | .................... | .................... | ..................... | .................... | .................... | .................... |
| V | 2638 | hCoV-19/Japan/PG-1768/2020\|EPI_ISL_685374\|2020-04 | .................... | .................... | ..................... | .................... | .................... | .................... |
| V | 2639 | hCoV-19/Japan/PG-1769/2020\|EPI_ISL_685375\|2020-04 | .................... | .................... | ..................... | .................... | .................... | .................... |
| V | 2640 | hCoV-19/Japan/PG-1770/2020\|EPI_ISL_685376\|2020-04 | .................... | .................... | ..................... | .................... | .................... | .................... |
| V | 2641 | hCoV-19/Japan/PG-1771/2020\|EPI_ISL_685377\|2020-04 | .................... | .................... | ..................... | .................... | .................... | .................... |
| V | 2642 | hCoV-19/Japan/PG-1772/2020\|EPI_ISL_685378\|2020-04 | .................... | .................... | ..................... | .................... | .................... | .................... |
| V | 2643 | hCoV-19/Japan/PG-1773/2020\|EPI_ISL_685379\|2020-04 | .................... | .................... | ..................... | .................... | .................... | .................... |
| V | 2644 | hCoV-19/Japan/PG-7597/2020\|EPI_ISL_686699\|2020-07 | .................... | .................... | ..................... | .................... | .................... | .................... |
| V | 2645 | hCoV-19/Japan/PG-7909/2020\|EPI_ISL_687010\|2020-08 | .................... | .................... | ..................... | .................... | .................... | .................... |
| V | 2646 | hCoV-19/Japan/PG-7910/2020\|EPI_ISL_687011\|2020-08 | .................... | .................... | ..................... | .................... | .................... | .................... |
| V | 2647 | hCoV-19/Japan/PG-7911/2020\|EPI_ISL_687012\|2020-08 | .................... | .................... | ..................... | .................... | .................... | .................... |
| V | 2648 | hCoV-19/Japan/PG-7912/2020\|EPI_ISL_687013\|2020-08 | .................... | .................... | ..................... | .................... | .................... | .................... |
| V | 2649 | hCoV-19/Japan/PG-7913/2020\|EPI_ISL_687014\|2020-08 | .................... | .................... | ..................... | .................... | .................... | .................... |
| V | 2650 | hCoV-19/Japan/PG-7914/2020\|EPI_ISL_687015\|2020-08 | .................... | .................... | ..................... | .................... | .................... | .................... |
| V | 2651 | hCoV-19/Japan/PG-7915/2020\|EPI_ISL_687016\|2020-07 | .................... | .................... | ..................... | .................... | .................... | .................... |
| V | 2652 | hCoV-19/Japan/PG-7916/2020\|EPI_ISL_687017\|2020-07 | .................... | .................... | ..................... | .................... | .................... | .................... |
| V | 2653 | hCoV-19/Japan/PG-7917/2020\|EPI_ISL_687018\|2020-07 | .................... | .................... | ..................... | .................... | .................... | .................... |
| V | 2654 | hCoV-19/Japan/PG-7918/2020\|EPI_ISL_687019\|2020-07 | .................... | .................... | ..................... | .................... | .................... | .................... |
| V | 2655 | hCoV-19/Japan/PG-7919/2020\|EPI_ISL_687020\|2020-07 | .................... | .................... | ..................... | .................... | .................... | .................... |
| V | 2656 | hCoV-19/Japan/PG-7920/2020\|EPI_ISL_687021\|2020-07 | .................... | .................... | ..................... | .................... | .................... | .................... |
| V | 2657 | hCoV-19/Japan/PG-7921/2020\|EPI_ISL_687022\|2020-07 | .................... | .................... | ..................... | .................... | .................... | .................... |
| V | 2658 | hCoV-19/Japan/PG-7922/2020\|EPI_ISL_687023\|2020-07 | .................... | .................... | ..................... | .................... | .................... | .................... |
| V | 2659 | hCoV-19/Japan/PG-7923/2020\|EPI_ISL_687024\|2020-07 | .................... | .................... | ..................... | .................... | .................... | .................... |
| V | 2660 | hCoV-19/Japan/PG-7924/2020\|EPI_ISL_687025\|2020-07 | .................... | .................... | ..................... | .................... | .................... | .................... |
| V | 2661 | hCoV-19/Japan/PG-7925/2020\|EPI_ISL_687026\|2020-07 | .................... | .................... | ..................... | .................... | .................... | .................... |
| V | 2662 | hCoV-19/Japan/PG-7926/2020\|EPI_ISL_687027\|2020-07 | .................... | .................... | ..................... | .................... | .................... | .................... |
| V | 2663 | hCoV-19/Japan/PG-7927/2020\|EPI_ISL_687028\|2020-07 | .................... | .................... | ..................... | .................... | .................... | .................... |
| V | 2664 | hCoV-19/Japan/PG-7236/2020\|EPI_ISL_688340\|2020-08 | .................... | .................... | ..................... | .................... | .................... | .................... |
| V | 2665 | hCoV-19/Japan/PG-7237/2020\|EPI_ISL_688341\|2020-08 | .................... | .................... | ..................... | .................... | .................... | .................... |
| V | 2666 | hCoV-19/Japan/PG-7238/2020\|EPI_ISL_688342\|2020-08 | .................... | .................... | ..................... | .................... | .................... | .................... |
| V | 2667 | hCoV-19/Japan/PG-7239/2020\|EPI_ISL_688343\|2020-08 | .................... | .................... | ..................... | .................... | .................... | .................... |
| V | 2668 | hCoV-19/Japan/PG-7241/2020\|EPI_ISL_688344\|2020-08 | .................... | .................... | ..................... | .................... | .................... | .................... |
| V | 2669 | hCoV-19/Japan/PG-7242/2020\|EPI_ISL_688345\|2020-08 | .................... | .................... | ..................... | .................... | .................... | .................... |
| V | 2670 | hCoV-19/Japan/PG-7243/2020\|EPI_ISL_688346\|2020-08 | .................... | .................... | ..................... | .................... | .................... | .................... |
| V | 2671 | hCoV-19/Japan/PG-7244/2020\|EPI_ISL_688347\|2020-08 | .................... | .................... | ..................... | .................... | .................... | .................... |
| V | 2672 | hCoV-19/Japan/PG-7245/2020\|EPI_ISL_688348\|2020-08 | .................... | .................... | ..................... | .................... | .................... | .................... |
| V | 2673 | hCoV-19/Japan/PG-7246/2020\|EPI_ISL_688349\|2020-08 | .................... | .................... | ..................... | .................... | .................... | .................... |
| V | 2674 | hCoV-19/Japan/PG-7247/2020\|EPI_ISL_688350\|2020-08 | .................... | .................... | ..................... | .................... | .................... | .................... |
| V | 2675 | hCoV-19/Japan/PG-7248/2020\|EPI_ISL_688351\|2020-08 | .................... | .................... | ..................... | .................... | .................... | .................... |
| V | 2676 | hCoV-19/Japan/PG-7249/2020\|EPI_ISL_688352\|2020-08 | .................... | .................... | ..................... | .................... | .................... | .................... |
| V | 2677 | hCoV-19/Japan/PG-7250/2020\|EPI_ISL_688353\|2020-08 | .................... | .................... | ..................... | .................... | .................... | .................... |
| V | 2678 | hCoV-19/Japan/PG-7251/2020\|EPI_ISL_688354\|2020-08 | .................... | .................... | ..................... | .................... | .................... | .................... |
| V | 2679 | hCoV-19/Japan/PG-7252/2020\|EPI_ISL_688355\|2020-08 | .................... | .................... | ..................... | .................... | .................... | .................... |
| V | 2680 | hCoV-19/Japan/PG-7253/2020\|EPI_ISL_688356\|2020-08 | .................... | .................... | ..................... | .................... | .................... | .................... |
| V | 2681 | hCoV-19/Japan/PG-7254/2020\|EPI_ISL_688357\|2020-08 | .................... | .................... | ..................... | .................... | .................... | .................... |
| V | 2682 | hCoV-19/Japan/PG-7255/2020\|EPI_ISL_688358\|2020-08 | .................... | .................... | ..................... | .................... | .................... | .................... |
| V | 2683 | hCoV-19/Japan/PG-7256/2020\|EPI_ISL_688359\|2020-08 | .................... | .................... | ..................... | .................... | .................... | .................... |
| V | 2684 | hCoV-19/Japan/PG-4532/2020\|EPI_ISL_689670\|2020-07 | .................... | .................... | ..................... | .................... | .................... | .................... |
| V | 2685 | hCoV-19/Japan/PG-4533/2020\|EPI_ISL_689671\|2020-07 | .................... | .................... | ..................... | .................... | .................... | .................... |
| V | 2686 | hCoV-19/Japan/PG-4534/2020\|EPI_ISL_689672\|2020-07 | .................... | .................... | ..................... | .................... | .................... | .................... |
| V | 2687 | hCoV-19/Japan/PG-4535/2020\|EPI_ISL_689673\|2020-07 | .................... | .................... | ..................... | .................... | .................... | .................... |
| V | 2688 | hCoV-19/Japan/PG-4536/2020\|EPI_ISL_689674\|2020-07 | .................... | .................... | ..................... | .................... | .................... | .................... |
| V | 2689 | hCoV-19/Japan/PG-4537/2020\|EPI_ISL_689675\|2020-07 | .................... | .................... | ..................... | .................... | .................... | .................... |
| V | 2690 | hCoV-19/Japan/PG-4538/2020\|EPI_ISL_689676\|2020-07 | .................... | .................... | ..................... | .................... | .................... | .................... |
| V | 2691 | hCoV-19/Japan/PG-4539/2020\|EPI_ISL_689677\|2020-07 | .................... | .................... | ..................... | .................... | .................... | .................... |
| V | 2692 | hCoV-19/Japan/PG-4540/2020\|EPI_ISL_689678\|2020-07 | .................... | .................... | ..................... | .................... | .................... | .................... |
| V | 2693 | hCoV-19/Japan/PG-4541/2020\|EPI_ISL_689679\|2020-07 | .................... | .................... | ..................... | .................... | .................... | .................... |
| V | 2694 | hCoV-19/Japan/PG-4542/2020\|EPI_ISL_689680\|2020 | .................... | .................... | ..................... | .................... | .................... | .................... |
| V | 2695 | hCoV-19/Japan/PG-4543/2020\|EPI_ISL_689681\|2020-07 | .................... | .................... | ..................... | .................... | .................... | .................... |

V 2696 hCoV-19/Japan/PG-4544/2020|EPI_ISL_689682|2020-07 .................... .................... ..................... .................... .................... ....................
V 2697 hCoV-19/Japan/PG-4545/2020|EPI_ISL_689683|2020-07 .................... .................... ..................... .................... .................... ....................
V 2698 hCoV-19/Japan/PG-4546/2020|EPI_ISL_689684|2020-07 .................... .................... ..................... .................... .................... ....................
V 2699 hCoV-19/Japan/PG-4547/2020|EPI_ISL_689685|2020-07 .................... .................... ..................... .................... .................... ....................
V 2701 hCoV-19/Japan/PG-4549/2020|EPI_ISL_689687|2020 .................... .................... ..................... .................... .................... ....................
V 2703 hCoV-19/Japan/PG-4551/2020|EPI_ISL_689689|2020 .................... .................... ..................... .................... .................... ....................
V 2704 hCoV-19/Japan/PG-4563/2020|EPI_ISL_689700|2020-07 .................... .................... ..................... .................... .................... ....................
V 2705 hCoV-19/Japan/PG-4564/2020|EPI_ISL_689701|2020-07 .................... .................... ..................... .................... .................... ....................
V 2706 hCoV-19/Japan/PG-4565/2020|EPI_ISL_689702|2020-07 .................... .................... ..................... .................... .................... ....................
V 2707 hCoV-19/Japan/PG-4566/2020|EPI_ISL_689703|2020-07 .................... .................... ..................... .................... .................... ....................
V 2708 hCoV-19/Japan/PG-4567/2020|EPI_ISL_689704|2020-07 .................... .................... ..................... .................... .................... ....................
V 2709 hCoV-19/Japan/PG-4568/2020|EPI_ISL_689705|2020-07 .................... .................... ..................... .................... .................... ....................
V 2710 hCoV-19/Japan/PG-4569/2020|EPI_ISL_689706|2020-07 .................... .................... ..................... .................... .................... ....................
V 2711 hCoV-19/Japan/PG-4570/2020|EPI_ISL_689707|2020-07 .................... .................... ..................... .................... .................... ....................
V 2712 hCoV-19/Japan/PG-4571/2020|EPI_ISL_689708|2020-07 .................... .................... ..................... .................... .................... ....................
V 2713 hCoV-19/Japan/PG-4572/2020|EPI_ISL_689709|2020-07 .................... .................... ..................... .................... .................... ....................
V 2714 hCoV-19/Japan/PG-4573/2020|EPI_ISL_689710|2020-07 .................... .................... ..................... .................... .................... ....................
V 2715 hCoV-19/Japan/PG-4574/2020|EPI_ISL_689711|2020-07 .................... .................... ..................... .................... .................... ....................
V 2716 hCoV-19/Japan/PG-4575/2020|EPI_ISL_689712|2020-07 .................... .................... ..................... .................... .................... ....................
V 2717 hCoV-19/Japan/PG-4576/2020|EPI_ISL_689713|2020-07 .................... .................... ..................... .................... .................... ....................
V 2718 hCoV-19/Japan/PG-4577/2020|EPI_ISL_689714|2020-07 .................... .................... ..................... .................... .................... ....................
V 2719 hCoV-19/Japan/PG-4578/2020|EPI_ISL_689715|2020-07 .................... .................... ..................... .................... .................... ....................
V 2720 hCoV-19/Japan/PG-4579/2020|EPI_ISL_689716|2020-07 .................... .................... ..................... .................... .................... ....................
V 2721 hCoV-19/Japan/PG-4580/2020|EPI_ISL_689717|2020-07 .................... .................... ..................... .................... .................... ....................
V 2722 hCoV-19/Japan/PG-4581/2020|EPI_ISL_689718|2020-07 .................... .................... ..................... .................... .................... ....................
V 2723 hCoV-19/Japan/PG-4582/2020|EPI_ISL_689719|2020-07 .................... .................... ..................... .................... .................... ....................
V 2724 hCoV-19/Japan/PG-4583/2020|EPI_ISL_689720|2020-08 .................... .................... ..................... .................... .................... ....................
V 2725 hCoV-19/Japan/PG-4584/2020|EPI_ISL_689721|2020-08 .................... .................... ..................... .................... .................... ....................
V 2726 hCoV-19/Japan/PG-4585/2020|EPI_ISL_689722|2020-08 .................... .................... ..................... .................... .................... ....................
V 2727 hCoV-19/Japan/PG-4586/2020|EPI_ISL_689723|2020-08 .................... .................... ..................... .................... .................... ....................
V 2728 hCoV-19/Japan/PG-2535/2020|EPI_ISL_690670|2020-05 .................... .................... ..................... .................... .................... ....................
V 2729 hCoV-19/Japan/PG-2536/2020|EPI_ISL_690671|2020-05 .................... .................... ..................... .................... .................... ....................
V 2730 hCoV-19/Japan/PG-2537/2020|EPI_ISL_690672|2020-05 .................... .................... ..................... .................... .................... ....................
V 2731 hCoV-19/Japan/PG-2538/2020|EPI_ISL_690673|2020-05 .................... .................... ..................... .................... .................... ....................
V 2732 hCoV-19/Japan/PG-2539/2020|EPI_ISL_690674|2020-05 .................... .................... ..................... .................... .................... ....................
V 2733 hCoV-19/Japan/PG-2540/2020|EPI_ISL_690675|2020-05 .................... .................... ..................... .................... .................... ....................
V 2734 hCoV-19/Japan/PG-2541/2020|EPI_ISL_690676|2020-05 .................... .................... ..................... .................... .................... ....................
V 2735 hCoV-19/Japan/PG-2542/2020|EPI_ISL_690677|2020-05 .................... .................... ..................... .................... .................... ....................
V 2736 hCoV-19/Japan/PG-2543/2020|EPI_ISL_690678|2020-05 .................... .................... ..................... .................... .................... ....................
V 2737 hCoV-19/Japan/PG-2544/2020|EPI_ISL_690679|2020-05 .................... .................... ..................... .................... .................... ....................
V 2738 hCoV-19/Japan/PG-2545/2020|EPI_ISL_690680|2020-05 .................... .................... ..................... .................... .................... ....................
V 2739 hCoV-19/Japan/PG-2546/2020|EPI_ISL_690681|2020-05 .................... .................... ..................... .................... .................... ....................
V 2740 hCoV-19/Japan/PG-2547/2020|EPI_ISL_690682|2020-05 .................... .................... ..................... .................... .................... ....................
V 2741 hCoV-19/Japan/PG-2548/2020|EPI_ISL_690683|2020-05 .................... .................... ..................... .................... .................... ....................
V 2742 hCoV-19/Japan/PG-2549/2020|EPI_ISL_690684|2020-04 .................... .................... ..................... .................... .................... ....................
V 2743 hCoV-19/Japan/PG-2550/2020|EPI_ISL_690685|2020-04 .................... .................... ..................... .................... .................... ....................
V 2744 hCoV-19/Japan/PG-2551/2020|EPI_ISL_690686|2020-04 .................... .................... ..................... .................... .................... ....................
V 2745 hCoV-19/Japan/PG-2554/2020|EPI_ISL_690689|2020-06 .................... .................... ..................... .................... .................... ....................
V 2746 hCoV-19/Japan/PG-2565/2020|EPI_ISL_690700|2020-06 .................... .................... ..................... .................... .................... ....................
V 2747 hCoV-19/Japan/PG-2566/2020|EPI_ISL_690701|2020-06 .................... .................... ..................... .................... .................... ....................
V 2748 hCoV-19/Japan/PG-2567/2020|EPI_ISL_690702|2020-06 .................... .................... ..................... .................... .................... ....................
V 2749 hCoV-19/Japan/PG-2568/2020|EPI_ISL_690703|2020-06 .................... .................... ..................... .................... .................... ....................
V 2750 hCoV-19/Japan/PG-2569/2020|EPI_ISL_690704|2020-06 .................... .................... ..................... .................... .................... ....................
V 2751 hCoV-19/Japan/PG-2571/2020|EPI_ISL_690705|2020-06 .................... .................... ..................... .................... .................... ....................
V 2752 hCoV-19/Japan/PG-2572/2020|EPI_ISL_690706|2020-06 .................... .................... ..................... .................... .................... ....................
V 2753 hCoV-19/Japan/PG-2573/2020|EPI_ISL_690707|2020-06 .................... .................... ..................... .................... .................... ....................
V 2754 hCoV-19/Japan/PG-2574/2020|EPI_ISL_690708|2020-03 .................... .................... ..................... .................... .................... ....................
V 2755 hCoV-19/Japan/PG-2575/2020|EPI_ISL_690709|2020-03 .................... .................... ..................... .................... .................... ....................
V 2756 hCoV-19/Japan/PG-3102/2020|EPI_ISL_691224|2020-04 .................... .................... ..................... .................... .................... ....................
V 2757 hCoV-19/Japan/PG-3103/2020|EPI_ISL_691225|2020-04 .................... .................... ..................... .................... .................... ....................
V 2758 hCoV-19/Japan/PG-3105/2020|EPI_ISL_691227|2020-05 .................... .................... ..................... .................... .................... ....................
V 2759 hCoV-19/Japan/PG-3108/2020|EPI_ISL_691230|2020-04 .................... .................... ..................... .................... .................... ....................
V 2760 hCoV-19/Japan/PG-3109/2020|EPI_ISL_691231|2020-04 .................... .................... ..................... .................... .................... ....................
V 2761 hCoV-19/Japan/PG-3110/2020|EPI_ISL_691232|2020-05 .................... .................... ..................... .................... .................... ....................
V 2762 hCoV-19/Japan/PG-3111/2020|EPI_ISL_691233|2020-05 .................... .................... ..................... .................... .................... ....................
V 2763 hCoV-19/Japan/PG-3112/2020|EPI_ISL_691234|2020-05 .................... .................... ..................... .................... .................... ....................
V 2764 hCoV-19/Japan/PG-3113/2020|EPI_ISL_691235|2020-05 .................... .................... ..................... .................... .................... ....................
V 2765 hCoV-19/Japan/PG-3114/2020|EPI_ISL_691236|2020-05 .................... .................... ..................... .................... .................... ....................
V 2766 hCoV-19/Japan/PG-3115/2020|EPI_ISL_691237|2020-05 .................... .................... ..................... .................... .................... ....................
V 2767 hCoV-19/Japan/PG-3116/2020|EPI_ISL_691238|2020-05 .................... .................... ..................... .................... .................... ....................
V 2768 hCoV-19/Japan/PG-3117/2020|EPI_ISL_691239|2020-05 .................... .................... ..................... .................... .................... ....................
V 2769 hCoV-19/Japan/PG-3118/2020|EPI_ISL_691240|2020-05 .................... .................... ..................... .................... .................... ....................
V 2770 hCoV-19/Japan/PG-3119/2020|EPI_ISL_691241|2020-05 .................... .................... ..................... .................... .................... ....................
V 2771 hCoV-19/Japan/PG-4327/2020|EPI_ISL_692555|2020-07 .................... .................... ..................... .................... .................... ....................
V 2772 hCoV-19/Japan/PG-4328/2020|EPI_ISL_692556|2020-07 .................... .................... ..................... .................... .................... ....................
V 2773 hCoV-19/Japan/PG-4332/2020|EPI_ISL_692560|2020-07 .................... .................... ..................... .................... .................... ....................
V 2774 hCoV-19/Japan/PG-4333/2020|EPI_ISL_692561|2020-07 .................... .................... ..................... .................... .................... ....................
V 2775 hCoV-19/Japan/PG-4334/2020|EPI_ISL_692562|2020-07 .................... .................... ..................... .................... .................... ....................
V 2776 hCoV-19/Japan/PG-4335/2020|EPI_ISL_692563|2020-07 .................... .................... ..................... .................... .................... ....................
V 2777 hCoV-19/Japan/PG-4336/2020|EPI_ISL_692564|2020-07 .................... .................... ..................... .................... .................... ....................

| | | | | | | | | |
|---|---|---|---|---|---|---|---|---|
| V | 2778 hCoV-19/Japan/PG-4337/2020\|EPI_ISL_692565\|2020-07 | .................... | .................... | .................... | .................... | .................... | .................... | |
| V | 2779 hCoV-19/Japan/PG-4338/2020\|EPI_ISL_692566\|2020-07 | .................... | .................... | .................... | .................... | .................... | .................... | |
| V | 2780 hCoV-19/Japan/PG-4339/2020\|EPI_ISL_692567\|2020-07 | .................... | .................... | .................... | .................... | .................... | .................... | |
| V | 2781 hCoV-19/Japan/PG-4340/2020\|EPI_ISL_692568\|2020-07 | .................... | .................... | .................... | .................... | .................... | .................... | |
| V | 2782 hCoV-19/Japan/PG-4341/2020\|EPI_ISL_692569\|2020-07 | .................... | .................... | .................... | .................... | .................... | .................... | |
| V | 2783 hCoV-19/Japan/PG-4342/2020\|EPI_ISL_692570\|2020-07 | .................... | .................... | .................... | .................... | .................... | .................... | |
| V | 2784 hCoV-19/Japan/PG-4343/2020\|EPI_ISL_692571\|2020-07 | .................... | .................... | .................... | .................... | .................... | .................... | |
| V | 2785 hCoV-19/Japan/PG-4344/2020\|EPI_ISL_692572\|2020-07 | .................... | .................... | .................... | .................... | .................... | .................... | |
| V | 2786 hCoV-19/Switzerland/GE-ETHZ-350100/2020\|EPI_ISL_69 | .................... | .................... | .................... | .................... | .................... | .................... | |
| V | 2787 hCoV-19/Switzerland/GR-ETHZ-350102/2020\|EPI_ISL_69 | .................... | .................... | .................... | ............R....... | .................Y.. | .................... | 1 |
| V | 2788 hCoV-19/Switzerland/SO-ETHZ-350103/2020\|EPI_ISL_69 | .................... | .................... | .................... | .................... | .................... | .................... | |
| V | 2789 hCoV-19/Switzerland/BE-ETHZ-350104/2020\|EPI_ISL_69 | .................... | .................... | .................... | .................... | .................... | .................... | |
| V | 2790 hCoV-19/Switzerland/GE-ETHZ-350105/2020\|EPI_ISL_69 | .................... | .................... | .................... | .................... | .................... | .................... | |
| V | 2791 hCoV-19/Switzerland/VD-ETHZ-350106/2020\|EPI_ISL_69 | .................... | .................... | .................... | .................... | .................... | .................... | |
| V | 2792 hCoV-19/Switzerland/SO-ETHZ-350107/2020\|EPI_ISL_69 | .................... | .A.................. | .................... | .................... | .................... | .................... | |
| V | 2793 hCoV-19/Switzerland/BE-ETHZ-350108/2020\|EPI_ISL_69 | .................... | .................... | .................... | .................... | ......K............. | .................... | |
| V | 2794 hCoV-19/Switzerland/SO-ETHZ-350109/2020\|EPI_ISL_69 | .................... | .................... | .................... | .................... | .................... | .................... | |
| V | 2795 hCoV-19/Switzerland/SZ-ETHZ-350110/2020\|EPI_ISL_69 | .................... | .................... | .................... | .................... | .................... | .................... | |
| V | 2796 hCoV-19/Switzerland/BE-ETHZ-350111/2020\|EPI_ISL_69 | .................... | .................... | .................... | .................... | .................... | .................... | |
| V | 2797 hCoV-19/Switzerland/AG-ETHZ-350112/2020\|EPI_ISL_69 | .................... | .................... | .................... | .................... | .................... | .................... | |
| V | 2798 hCoV-19/Switzerland/AG-ETHZ-350113/2020\|EPI_ISL_69 | .................... | .................... | .................... | .................... | .................... | .................... | |
| V | 2799 hCoV-19/Switzerland/ZH-ETHZ-350114/2020\|EPI_ISL_69 | .................... | .................... | .................... | .................... | .................... | .................... | |
| V | 2800 hCoV-19/Switzerland/ZH-ETHZ-350115/2020\|EPI_ISL_69 | .................... | .................... | .................... | .................... | .................... | .................... | |
| V | 2801 hCoV-19/Switzerland/VS-ETHZ-350116/2020\|EPI_ISL_69 | .................... | .................... | .................... | .................... | .................... | .................... | |
| V | 2802 hCoV-19/Switzerland/AG-ETHZ-350117/2020\|EPI_ISL_69 | .................... | .................... | .................... | .................... | .................... | .................... | |
| V | 2803 hCoV-19/Switzerland/ZH-ETHZ-350119/2020\|EPI_ISL_69 | .................... | .................... | .................... | .................... | .................... | .................... | |
| V | 2804 hCoV-19/Switzerland/LU-ETHZ-350120/2020\|EPI_ISL_69 | .................... | .................... | .................... | .................... | ......K............. | .................... | |
| V | 2805 hCoV-19/Switzerland/VD-ETHZ-360048/2020\|EPI_ISL_69 | .................... | .................... | .................... | .................... | .................... | .................... | |
| V | 2806 hCoV-19/Switzerland/GR-ETHZ-360055/2020\|EPI_ISL_69 | .................... | .................... | .................... | .................... | .................... | .................... | |
| V | 2807 hCoV-19/Switzerland/BS-ETHZ-360056/2020\|EPI_ISL_69 | .................... | .................... | .................... | .................... | .................... | .................... | |
| V | 2808 hCoV-19/Switzerland/GE-ETHZ-360077/2020\|EPI_ISL_69 | .................... | .................... | .................... | .................... | .................... | .................... | |
| V | 2809 hCoV-19/Switzerland/VS-ETHZ-360078/2020\|EPI_ISL_69 | .................... | .................... | .................... | .................... | .................... | .................... | |
| V | 2810 hCoV-19/Switzerland/AG-ETHZ-360081/2020\|EPI_ISL_69 | .................... | .................... | .................... | .................... | .................... | .................... | |
| V | 2811 hCoV-19/Switzerland/SH-ETHZ-360083/2020\|EPI_ISL_69 | .................... | .................... | .................... | .................... | .................... | .................... | |
| V | 2812 hCoV-19/Switzerland/SH-ETHZ-360084/2020\|EPI_ISL_69 | .................... | .................... | .................... | .................... | .................... | .................... | |
| V | 2813 hCoV-19/Switzerland/BE-ETHZ-360086/2020\|EPI_ISL_69 | .................... | .................... | .................... | .................... | .................... | .................... | |
| V | 2814 hCoV-19/Switzerland/BE-ETHZ-360087/2020\|EPI_ISL_69 | .................... | .................... | .................... | .................... | .................... | .................... | |
| V | 2815 hCoV-19/Switzerland/BE-ETHZ-360088/2020\|EPI_ISL_69 | .................... | .................... | .................... | .................... | .................... | .................... | |
| V | 2816 hCoV-19/Switzerland/BE-ETHZ-360089/2020\|EPI_ISL_69 | .................... | .................... | .................... | .................... | .................... | .................... | |
| V | 2817 hCoV-19/Japan/PG-1190/2020\|EPI_ISL_684800\|2020-04 | .................... | .................... | .................... | .................... | .................... | .................... | |
| V | 2818 hCoV-19/Japan/PG-1191/2020\|EPI_ISL_684801\|2020-04 | .................... | .................... | .................... | .................... | .................... | .................... | |
| V | 2819 hCoV-19/Japan/PG-1192/2020\|EPI_ISL_684802\|2020-04 | .................... | .................... | .................... | .................... | .................... | .................... | |
| V | 2820 hCoV-19/Japan/PG-1193/2020\|EPI_ISL_684803\|2020-04 | .................... | .................... | .................... | .................... | .................... | .................... | |
| V | 2821 hCoV-19/Japan/PG-1194/2020\|EPI_ISL_684804\|2020-04 | .................... | .................... | .................... | .................... | .................... | .................... | |
| V | 2822 hCoV-19/Japan/PG-1195/2020\|EPI_ISL_684805\|2020-04 | .................... | .................... | .................... | .................... | .................... | .................... | |
| V | 2823 hCoV-19/Japan/PG-1196/2020\|EPI_ISL_684806\|2020-04 | .................... | .................... | .................... | .................... | .................... | .................... | |
| V | 2824 hCoV-19/Japan/PG-1197/2020\|EPI_ISL_684807\|2020-04 | .................... | .................... | .................... | .................... | .................... | .................... | |
| V | 2825 hCoV-19/Japan/PG-1198/2020\|EPI_ISL_684808\|2020-04 | .................... | .................... | .................... | .................... | .................... | .................... | |
| V | 2826 hCoV-19/Japan/PG-1199/2020\|EPI_ISL_684809\|2020-04 | .................... | .................... | .................... | .................... | .................... | .................... | |
| V | 2827 hCoV-19/Japan/PG-1200/2020\|EPI_ISL_684810\|2020-04 | .................... | .................... | .................... | .................... | .................... | .................... | |
| V | 2828 hCoV-19/Japan/PG-1201/2020\|EPI_ISL_684811\|2020-04 | .................... | .................... | .................... | .................... | .................... | .................... | |
| V | 2829 hCoV-19/Japan/PG-1202/2020\|EPI_ISL_684812\|2020-04 | .................... | .................... | .................... | .................... | .................... | .................... | |
| V | 2830 hCoV-19/Japan/PG-1203/2020\|EPI_ISL_684813\|2020-04 | .................... | .................... | .................... | .................... | .................... | .................... | |
| V | 2831 hCoV-19/Japan/PG-1205/2020\|EPI_ISL_684815\|2020-04 | .................... | .................... | .................... | .................... | .................... | .................... | |
| V | 2832 hCoV-19/Japan/PG-1206/2020\|EPI_ISL_684816\|2020-04 | .................... | .................... | .................... | .................... | .................... | .................... | |
| V | 2833 hCoV-19/Japan/PG-1207/2020\|EPI_ISL_684817\|2020-04 | .................... | .................... | .................... | .................... | .................... | .................... | |
| V | 2834 hCoV-19/Japan/PG-1208/2020\|EPI_ISL_684818\|2020-04 | .................... | .................... | .................... | .................... | .................... | .................... | |
| V | 2835 hCoV-19/Japan/PG-1209/2020\|EPI_ISL_684819\|2020-04 | .................... | .................... | .................... | .................... | .................... | .................... | |
| V | 2836 hCoV-19/Japan/PG-1210/2020\|EPI_ISL_684820\|2020-04 | .................... | .................... | .................... | .................... | .................... | .................... | |
| V | 2837 hCoV-19/Japan/PG-1211/2020\|EPI_ISL_684821\|2020-04 | .................... | .................... | .................... | .................... | .................... | .................... | |
| V | 2838 hCoV-19/Japan/PG-1212/2020\|EPI_ISL_684822\|2020-04 | .................... | .................... | .................... | .................... | .................... | .................... | |
| V | 2839 hCoV-19/Japan/PG-1213/2020\|EPI_ISL_684823\|2020-04 | .................... | .................... | .................... | .................... | .................... | .................... | |
| V | 2840 hCoV-19/Japan/PG-1214/2020\|EPI_ISL_684824\|2020-04 | .................... | .................... | .................... | .................... | .................... | .................... | |
| V | 2841 hCoV-19/Japan/PG-1215/2020\|EPI_ISL_684825\|2020-04 | .................... | .................... | .................... | .................... | .................... | .................... | |
| V | 2842 hCoV-19/Japan/PG-1216/2020\|EPI_ISL_684826\|2020-04 | .................... | .................... | .................... | .................... | .................... | .................... | |
| V | 2843 hCoV-19/Japan/PG-1217/2020\|EPI_ISL_684827\|2020-04 | .................... | .................... | .................... | .................... | .................... | .................... | |
| V | 2844 hCoV-19/Japan/PG-1218/2020\|EPI_ISL_684828\|2020-04 | .................... | .................... | .................... | .................... | .................... | .................... | |
| V | 2845 hCoV-19/Japan/PG-1219/2020\|EPI_ISL_684829\|2020-04 | .................... | .................... | .................... | .................... | .................... | .................... | |
| V | 2846 hCoV-19/Japan/PG-7899/2020\|EPI_ISL_687000\|2020-07 | .................... | .................... | .................... | .................... | .................... | .................... | |
| V | 2847 hCoV-19/Japan/PG-7900/2020\|EPI_ISL_687001\|2020-07 | .................... | .................... | .................... | .................... | .................... | .................... | |
| V | 2848 hCoV-19/Japan/PG-7901/2020\|EPI_ISL_687002\|2020-07 | .................... | .................... | .................... | .................... | .................... | .................... | |
| V | 2849 hCoV-19/Japan/PG-7902/2020\|EPI_ISL_687003\|2020-07 | .................... | .................... | .................... | .................... | .................... | .................... | |
| V | 2850 hCoV-19/Japan/PG-7903/2020\|EPI_ISL_687004\|2020-07 | .................... | .................... | .................... | .................... | .................... | .................... | |
| V | 2851 hCoV-19/Japan/PG-7904/2020\|EPI_ISL_687005\|2020-07 | .................... | .................... | .................... | .................... | .................... | .................... | |
| V | 2852 hCoV-19/Japan/PG-7905/2020\|EPI_ISL_687006\|2020-07 | .................... | .................... | .................... | .................... | .................... | .................... | |
| V | 2853 hCoV-19/Japan/PG-7906/2020\|EPI_ISL_687007\|2020-08 | .................... | .................... | .................... | .................... | .................... | .................... | |
| V | 2854 hCoV-19/Japan/PG-7907/2020\|EPI_ISL_687008\|2020-08 | .................... | .................... | .................... | .................... | .................... | .................... | |
| V | 2855 hCoV-19/Japan/PG-7908/2020\|EPI_ISL_687009\|2020-08 | .................... | .................... | .................... | .................... | .................... | .................... | |
| V | 2856 hCoV-19/Japan/PG-7962/2020\|EPI_ISL_687063\|2020-05 | .................... | .................... | .................... | .................... | .................... | .................... | |
| V | 2857 hCoV-19/Japan/PG-7963/2020\|EPI_ISL_687064\|2020-05 | .................... | .................... | .................... | .................... | .................... | .................... | |

| | | | | | | | | |
|---|---|---|---|---|---|---|---|---|
| V | 2858 | hCoV-19/Japan/PG-7964/2020\|EPI_ISL_687065\|2020-05 | .................... | .................... | ..................... | .................... | .................... | .................... |
| V | 2859 | hCoV-19/Japan/PG-7965/2020\|EPI_ISL_687066\|2020-05 | .................... | .................... | ..................... | .................... | .................... | .................... |
| V | 2860 | hCoV-19/Japan/PG-7966/2020\|EPI_ISL_687067\|2020-05 | .................... | .................... | ..................... | .................... | .................... | .................... |
| V | 2861 | hCoV-19/Japan/PG-7967/2020\|EPI_ISL_687068\|2020-05 | .................... | .................... | ..................... | .................... | .................... | .................... |
| V | 2862 | hCoV-19/Japan/PG-7968/2020\|EPI_ISL_687069\|2020-05 | .................... | .................... | ..................... | .................... | .................... | .................... |
| V | 2863 | hCoV-19/Japan/PG-7969/2020\|EPI_ISL_687070\|2020-05 | .................... | .................... | ..................... | .................... | .................... | .................... |
| V | 2864 | hCoV-19/Japan/PG-7970/2020\|EPI_ISL_687071\|2020-05 | .................... | .................... | ..................... | .................... | .................... | .................... |
| V | 2865 | hCoV-19/Japan/PG-7971/2020\|EPI_ISL_687072\|2020-05 | .................... | .................... | ..................... | .................... | .................... | .................... |
| V | 2866 | hCoV-19/Japan/PG-7972/2020\|EPI_ISL_687073\|2020-05 | .................... | .................... | ..................... | .................... | .................... | .................... |
| V | 2867 | hCoV-19/Japan/PG-7973/2020\|EPI_ISL_687074\|2020-07 | .................... | .................... | ..................... | .................... | .................... | .................... |
| V | 2868 | hCoV-19/Japan/PG-7974/2020\|EPI_ISL_687075\|2020-08 | .................... | .................... | ..................... | .................... | .................... | .................... |
| V | 2869 | hCoV-19/Japan/PG-7975/2020\|EPI_ISL_687076\|2020-08 | .................... | .................... | ..................... | .................... | .................... | .................... |
| V | 2870 | hCoV-19/Japan/PG-7976/2020\|EPI_ISL_687077\|2020-08 | .................... | .................... | ..................... | .................... | .................... | .................... |
| V | 2871 | hCoV-19/Japan/PG-7977/2020\|EPI_ISL_687078\|2020-08 | .................... | .A.................. | ..................... | .................... | .................... | .................... |
| V | 2872 | hCoV-19/Japan/PG-7978/2020\|EPI_ISL_687079\|2020-08 | .................... | .................... | ..................... | .................... | .................... | .................... |
| V | 2873 | hCoV-19/Japan/PG-7979/2020\|EPI_ISL_687080\|2020-08 | .................... | .................... | ..................... | .................... | .................... | .................... |
| V | 2874 | hCoV-19/Japan/PG-7980/2020\|EPI_ISL_687081\|2020-08 | .................... | .................... | ..................... | .................... | .................... | .................... |
| V | 2875 | hCoV-19/Japan/PG-7981/2020\|EPI_ISL_687082\|2020-08 | .................... | .................... | ..................... | .................... | .................... | .................... |
| V | 2876 | hCoV-19/Japan/PG-7982/2020\|EPI_ISL_687083\|2020-08 | .................... | .................... | ..................... | .................... | .................... | .................... |
| V | 2877 | hCoV-19/Japan/PG-7983/2020\|EPI_ISL_687084\|2020-08 | .................... | .................... | ..................... | .................... | .................... | .................... |
| V | 2878 | hCoV-19/Japan/PG-7984/2020\|EPI_ISL_687085\|2020-08 | .................... | .................... | ..................... | .................... | .................... | .................... |
| V | 2879 | hCoV-19/Japan/PG-7985/2020\|EPI_ISL_687086\|2020-08 | .................... | .................... | ..................... | .................... | .................... | .................... |
| V | 2880 | hCoV-19/Japan/PG-7986/2020\|EPI_ISL_687087\|2020-08 | .................... | .................... | ..................... | .................... | .................... | .................... |
| V | 2881 | hCoV-19/Japan/PG-7987/2020\|EPI_ISL_687088\|2020-08 | .................... | .................... | ..................... | .................... | .................... | .................... |
| V | 2882 | hCoV-19/Japan/PG-7988/2020\|EPI_ISL_687089\|2020-08 | .................... | .................... | ..................... | .................... | .................... | .................... |
| V | 2883 | hCoV-19/Japan/PG-7989/2020\|EPI_ISL_687090\|2020-08 | .................... | .................... | ..................... | .................... | .................... | .................... |
| V | 2884 | hCoV-19/Japan/PG-7990/2020\|EPI_ISL_687091\|2020-08 | .................... | .................... | ..................... | .................... | .................... | .................... |
| V | 2885 | hCoV-19/Japan/PG-7991/2020\|EPI_ISL_687092\|2020-08 | .................... | .................... | ..................... | .................... | .................... | .................... |
| V | 2886 | hCoV-19/Japan/PG-7992/2020\|EPI_ISL_687093\|2020-08 | .................... | .................... | ..................... | .................... | .................... | .................... |
| V | 2887 | hCoV-19/Japan/PG-7993/2020\|EPI_ISL_687094\|2020-08 | .................... | .................... | ..................... | .................... | .................... | .................... |
| V | 2888 | hCoV-19/Japan/PG-7994/2020\|EPI_ISL_687095\|2020-08 | .................... | .................... | ..................... | .................... | .................... | .................... |
| V | 2889 | hCoV-19/Japan/PG-7995/2020\|EPI_ISL_687096\|2020-08 | .................... | .................... | ..................... | .................... | .................... | .................... |
| V | 2890 | hCoV-19/Japan/PG-7996/2020\|EPI_ISL_687097\|2020-08 | .................... | .................... | ..................... | .................... | .................... | .................... |
| V | 2891 | hCoV-19/Japan/PG-7997/2020\|EPI_ISL_687098\|2020-08 | .................... | .................... | ..................... | .................... | .................... | .................... |
| V | 2892 | hCoV-19/Japan/PG-7998/2020\|EPI_ISL_687099\|2020-09 | .................... | .................... | ..................... | .................... | .................... | .................... |
| V | 2893 | hCoV-19/Japan/PG-7196/2020\|EPI_ISL_688300\|2020-07 | .................... | .................... | ..................... | .................... | .................... | .................... |
| V | 2894 | hCoV-19/Japan/PG-7197/2020\|EPI_ISL_688301\|2020-07 | .................... | .................... | ..................... | .................... | .................... | .................... |
| V | 2895 | hCoV-19/Japan/PG-7198/2020\|EPI_ISL_688302\|2020-07 | .................... | .................... | ..................... | .................... | .................... | .................... |
| V | 2896 | hCoV-19/Japan/PG-7199/2020\|EPI_ISL_688303\|2020-07 | .................... | .................... | ..................... | .................... | .................... | .................... |
| V | 2897 | hCoV-19/Japan/PG-7200/2020\|EPI_ISL_688304\|2020-07 | .................... | .................... | ..................... | .................... | .................... | .................... |
| V | 2898 | hCoV-19/Japan/PG-7201/2020\|EPI_ISL_688305\|2020-07 | .................... | .................... | ..................... | .................... | .................... | .................... |
| V | 2899 | hCoV-19/Japan/PG-7202/2020\|EPI_ISL_688306\|2020-07 | .................... | .................... | ..................... | .................... | .................... | .................... |
| V | 2900 | hCoV-19/Japan/PG-7203/2020\|EPI_ISL_688307\|2020-07 | .................... | .................... | ..................... | .................... | .................... | .................... |
| V | 2901 | hCoV-19/Japan/PG-7204/2020\|EPI_ISL_688308\|2020-07 | .................... | .................... | ..................... | .................... | .................... | .................... |
| V | 2902 | hCoV-19/Japan/PG-7205/2020\|EPI_ISL_688309\|2020-07 | .................... | .................... | ..................... | .................... | .................... | .................... |
| V | 2903 | hCoV-19/Japan/PG-7206/2020\|EPI_ISL_688310\|2020-07 | .................... | .................... | ..................... | .................... | .................... | .................... |
| V | 2904 | hCoV-19/Japan/PG-7207/2020\|EPI_ISL_688311\|2020-07 | .................... | .................... | ..................... | .................... | .................... | .................... |
| V | 2905 | hCoV-19/Japan/PG-7208/2020\|EPI_ISL_688312\|2020-07 | .................... | .................... | ..................... | .................... | .................... | .................... |
| V | 2906 | hCoV-19/Japan/PG-7209/2020\|EPI_ISL_688313\|2020-07 | .................... | .................... | ..................... | .................... | .................... | .................... |
| V | 2907 | hCoV-19/Japan/PG-7210/2020\|EPI_ISL_688314\|2020-07 | .................... | .................... | ..................... | .................... | .................... | .................... |
| V | 2908 | hCoV-19/Japan/PG-7211/2020\|EPI_ISL_688315\|2020-07 | .................... | .................... | ..................... | .................... | .................... | .................... |
| V | 2909 | hCoV-19/Japan/PG-7212/2020\|EPI_ISL_688316\|2020-07 | .................... | .................... | ..................... | .................... | .................... | .................... |
| V | 2910 | hCoV-19/Japan/PG-7213/2020\|EPI_ISL_688317\|2020-07 | .................... | .................... | ..................... | .................... | .................... | .................... |
| V | 2911 | hCoV-19/Japan/PG-7214/2020\|EPI_ISL_688318\|2020-07 | .................... | .................... | ..................... | .................... | .................... | .................... |
| V | 2912 | hCoV-19/Japan/PG-7215/2020\|EPI_ISL_688319\|2020-07 | .................... | .................... | ..................... | ............A....... | .................... | .................... |
| V | 2913 | hCoV-19/Japan/PG-7216/2020\|EPI_ISL_688320\|2020-07 | .................... | .................... | ..................... | .................... | .................... | .................... |
| V | 2914 | hCoV-19/Japan/PG-7217/2020\|EPI_ISL_688321\|2020-07 | .................... | .................... | ..................... | .................... | .................... | .................... |
| V | 2915 | hCoV-19/Japan/PG-7218/2020\|EPI_ISL_688322\|2020-07 | .................... | .................... | ..................... | .................... | .................... | .................... |
| V | 2916 | hCoV-19/Japan/PG-7219/2020\|EPI_ISL_688323\|2020-07 | .................... | .................... | ..................... | .................... | .................... | .................... |
| V | 2917 | hCoV-19/Japan/PG-7220/2020\|EPI_ISL_688324\|2020-07 | .................... | .................... | ..................... | .................... | .................... | .................... |
| V | 2918 | hCoV-19/Japan/PG-7221/2020\|EPI_ISL_688325\|2020-07 | .................... | .................... | ..................... | .................... | .................... | .................... |
| V | 2919 | hCoV-19/Japan/PG-7222/2020\|EPI_ISL_688326\|2020-07 | .................... | .................... | ..................... | .................... | .................... | .................... |
| V | 2920 | hCoV-19/Japan/PG-7223/2020\|EPI_ISL_688327\|2020-08 | .................... | .................... | ..................... | .................... | .................... | .................... |
| V | 2921 | hCoV-19/Japan/PG-7224/2020\|EPI_ISL_688328\|2020-08 | .................... | .................... | ..................... | .................... | .................... | .................... |
| V | 2922 | hCoV-19/Japan/PG-7225/2020\|EPI_ISL_688329\|2020-08 | .................... | .................... | ..................... | .................... | .................... | .................... |
| V | 2923 | hCoV-19/Japan/PG-7226/2020\|EPI_ISL_688330\|2020-08 | .................... | .................... | ..................... | .................... | .................... | .................... |
| V | 2924 | hCoV-19/Japan/PG-7227/2020\|EPI_ISL_688331\|2020-07 | .................... | .................... | ..................... | .................... | .................... | .................... |
| V | 2925 | hCoV-19/Japan/PG-7228/2020\|EPI_ISL_688332\|2020-07 | .................... | .................... | ..................... | .................... | .................... | .................... |
| V | 2926 | hCoV-19/Japan/PG-7229/2020\|EPI_ISL_688333\|2020-07 | .................... | .................... | ..................... | .................... | .................... | .................... |
| V | 2927 | hCoV-19/Japan/PG-7230/2020\|EPI_ISL_688334\|2020-08 | .................... | .................... | ..................... | .................... | .................... | .................... |
| V | 2928 | hCoV-19/Japan/PG-7231/2020\|EPI_ISL_688335\|2020-08 | .................... | .................... | ..................... | .................... | .................... | .................... |
| V | 2929 | hCoV-19/Japan/PG-7232/2020\|EPI_ISL_688336\|2020-08 | .................... | .................... | ..................... | .................... | .................... | .................... |
| V | 2930 | hCoV-19/Japan/PG-7233/2020\|EPI_ISL_688337\|2020-08 | .................... | .................... | ..................... | .................... | .................... | .................... |
| V | 2931 | hCoV-19/Japan/PG-7234/2020\|EPI_ISL_688338\|2020-08 | .................... | .................... | ..................... | .................... | .................... | .................... |
| V | 2932 | hCoV-19/Japan/PG-7235/2020\|EPI_ISL_688339\|2020-08 | .................... | .................... | ..................... | .................... | .................... | .................... |
| V | 2933 | hCoV-19/Japan/PG-7290/2020\|EPI_ISL_688393\|2020-08 | .................... | .................... | ..................... | .................... | .................... | .................... |
| V | 2934 | hCoV-19/Japan/PG-7291/2020\|EPI_ISL_688394\|2020-08 | .................... | .................... | ..................... | .................... | .................... | .................... |
| V | 2935 | hCoV-19/Japan/PG-7292/2020\|EPI_ISL_688395\|2020-08 | .................... | .................... | ..................... | .................... | .................... | .................... |
| V | 2936 | hCoV-19/Japan/PG-7293/2020\|EPI_ISL_688396\|2020-08 | .................... | .................... | ..................... | .................... | .................... | .................... |
| V | 2937 | hCoV-19/Japan/PG-7294/2020\|EPI_ISL_688397\|2020-08 | .................... | .................... | ..................... | .................... | .................... | .................... |

| | | | | | | | | |
|---|---|---|---|---|---|---|---|---|
| V | 2938 | hCoV-19/Japan/PG-7295/2020\|EPI_ISL_688398\|2020-08 | .................... | .................... | .................... | .................... | .................... | .................... |
| V | 2939 | hCoV-19/Japan/PG-7296/2020\|EPI_ISL_688399\|2020-08 | .................... | .................... | .................... | .................... | .................... | .................... |
| V | 2940 | hCoV-19/Japan/PG-6501/2020\|EPI_ISL_689603\|2020-07 | .................... | .................... | .................... | .................... | .................... | .................... |
| V | 2941 | hCoV-19/Japan/PG-6502/2020\|EPI_ISL_689604\|2020-07 | .................... | .................... | .................... | .................... | .................... | .................... |
| V | 2942 | hCoV-19/Japan/PG-6503/2020\|EPI_ISL_689605\|2020-07 | .................... | .................... | .................... | .................... | .................... | .................... |
| V | 2943 | hCoV-19/Japan/PG-6504/2020\|EPI_ISL_689606\|2020-07 | .................... | .................... | .................... | .................... | .................... | .................... |
| V | 2944 | hCoV-19/Japan/PG-6505/2020\|EPI_ISL_689607\|2020-07 | .................... | .................... | .................... | .................... | .................... | .................... |
| V | 2945 | hCoV-19/Japan/PG-6506/2020\|EPI_ISL_689608\|2020-07 | .................... | .................... | .................... | .................... | .................... | .................... |
| V | 2946 | hCoV-19/Japan/PG-6507/2020\|EPI_ISL_689609\|2020-07 | .................... | .................... | .................... | .................... | .................... | .................... |
| V | 2947 | hCoV-19/Japan/PG-6508/2020\|EPI_ISL_689610\|2020-07 | .................... | .................... | .................... | .................... | .................... | .................... |
| V | 2948 | hCoV-19/Japan/PG-6509/2020\|EPI_ISL_689611\|2020-07 | .................... | .................... | .................... | .................... | .................... | .................... |
| V | 2949 | hCoV-19/Japan/PG-4714/2020\|EPI_ISL_689624\|2020-07 | .................... | .................... | .................... | .................... | .................... | .................... |
| V | 2950 | hCoV-19/Japan/PG-4778/2020\|EPI_ISL_689625\|2020-08 | .................... | .................... | .................... | .................... | .................... | .................... |
| V | 2951 | hCoV-19/Japan/PG-4779/2020\|EPI_ISL_689626\|2020-08 | .................... | .................... | .................... | .................... | .................... | .................... |
| V | 2952 | hCoV-19/Japan/PG-4780/2020\|EPI_ISL_689627\|2020-08 | .................... | .................... | .................... | .................... | .................... | .................... |
| V | 2953 | hCoV-19/Japan/PG-4781/2020\|EPI_ISL_689628\|2020-08 | .................... | .................... | .................... | .................... | .................... | .................... |
| V | 2954 | hCoV-19/Japan/PG-4782/2020\|EPI_ISL_689629\|2020-08 | .................... | .................... | .................... | .................... | .................... | .................... |
| V | 2955 | hCoV-19/Japan/PG-4783/2020\|EPI_ISL_689630\|2020-08 | .................... | .................... | .................... | .................... | .................... | .................... |
| V | 2956 | hCoV-19/Japan/PG-4784/2020\|EPI_ISL_689631\|2020-08 | .................... | .................... | .................... | .................... | .................... | .................... |
| V | 2957 | hCoV-19/Japan/PG-4789/2020\|EPI_ISL_689632\|2020-08 | .................... | .................... | .................... | .................... | .................... | .................... |
| V | 2958 | hCoV-19/Japan/PG-4790/2020\|EPI_ISL_689633\|2020-08 | .................... | .................... | .................... | .................... | .................... | .................... |
| V | 2959 | hCoV-19/Japan/PG-4792/2020\|EPI_ISL_689634\|2020-08 | .................... | .................... | .................... | .................... | .................... | .................... |
| V | 2960 | hCoV-19/Japan/PG-4982/2020\|EPI_ISL_689635\|2020-08 | .................... | .................... | .................... | .................... | .................... | .................... |
| V | 2961 | hCoV-19/Japan/PG-4983/2020\|EPI_ISL_689636\|2020-08 | .................... | .................... | .................... | .................... | .................... | .................... |
| V | 2962 | hCoV-19/Japan/PG-4985/2020\|EPI_ISL_689637\|2020-08 | .................... | .................... | .................... | .................... | .................... | .................... |
| V | 2963 | hCoV-19/Japan/PG-5007/2020\|EPI_ISL_689638\|2020-08 | .................... | .................... | .................... | .................... | .................... | .................... |
| V | 2964 | hCoV-19/Japan/PG-5011/2020\|EPI_ISL_689639\|2020-08 | .................... | .................... | .................... | .................... | .................... | .................... |
| V | 2965 | hCoV-19/Japan/PG-4838/2020\|EPI_ISL_689641\|2020-07 | .................... | .................... | .................... | .................... | .................... | .................... |
| V | 2966 | hCoV-19/Japan/PG-4938/2020\|EPI_ISL_689642\|2020-07 | .................... | .................... | .................... | .................... | .................... | .................... |
| V | 2967 | hCoV-19/Japan/PG-5100/2020\|EPI_ISL_689643\|2020 | .................... | .................... | .................... | .................... | .................... | .................... |
| V | 2968 | hCoV-19/Japan/PG-5161/2020\|EPI_ISL_689644\|2020-08 | .................... | .................... | .................... | .................... | .................... | .................... |
| V | 2969 | hCoV-19/Japan/PG-5483/2020\|EPI_ISL_689645\|2020-08 | .................... | .................... | .................... | .................... | .................... | .................... |
| V | 2970 | hCoV-19/Japan/PG-4510/2020\|EPI_ISL_689649\|2020-07 | .................... | .................... | .................... | .................... | .................... | .................... |
| V | 2971 | hCoV-19/Japan/PG-4511/2020\|EPI_ISL_689650\|2020-07 | .................... | .................... | .................... | .................... | .................... | .................... |
| V | 2972 | hCoV-19/Japan/PG-4512/2020\|EPI_ISL_689651\|2020-07 | .................... | .................... | .................... | .................... | .................... | .................... |
| V | 2973 | hCoV-19/Japan/PG-4513/2020\|EPI_ISL_689652\|2020-07 | .................... | .................... | .................... | .................... | .................... | .................... |
| V | 2974 | hCoV-19/Japan/PG-4514/2020\|EPI_ISL_689653\|2020-07 | .................... | .................... | .................... | .................... | .................... | .................... |
| V | 2975 | hCoV-19/Japan/PG-4515/2020\|EPI_ISL_689654\|2020-07 | .................... | .................... | .................... | .................... | .................... | .................... |
| V | 2976 | hCoV-19/Japan/PG-4516/2020\|EPI_ISL_689655\|2020-07 | .................... | .................... | .................... | .................... | .................... | .................... |
| V | 2977 | hCoV-19/Japan/PG-4517/2020\|EPI_ISL_689656\|2020-07 | .................... | .................... | .................... | .................... | .................... | .................... |
| V | 2978 | hCoV-19/Japan/PG-4518/2020\|EPI_ISL_689657\|2020-07 | .................... | .................... | .................... | .................... | .................... | .................... |
| V | 2979 | hCoV-19/Japan/PG-4519/2020\|EPI_ISL_689658\|2020-07 | .................... | .................... | .................... | .................... | .................... | .................... |
| V | 2980 | hCoV-19/Japan/PG-4520/2020\|EPI_ISL_689659\|2020-07 | .................... | .................... | .................... | .................... | .................... | .................... |
| V | 2981 | hCoV-19/Japan/PG-4521/2020\|EPI_ISL_689660\|2020-07 | .................... | .................... | .................... | .................... | .................... | .................... |
| V | 2982 | hCoV-19/Japan/PG-4522/2020\|EPI_ISL_689661\|2020-08 | .................... | .................... | .................... | .................... | .................... | .................... |
| V | 2983 | hCoV-19/Japan/PG-4523/2020\|EPI_ISL_689662\|2020-08 | .................... | .................... | .................... | .................... | .................... | .................... |
| V | 2984 | hCoV-19/Japan/PG-4524/2020\|EPI_ISL_689663\|2020-08 | .................... | .................... | .................... | .................... | .................... | .................... |
| V | 2985 | hCoV-19/Japan/PG-4526/2020\|EPI_ISL_689664\|2020-07 | .................... | .................... | .................... | .................... | .................... | .................... |
| V | 2986 | hCoV-19/Japan/PG-4527/2020\|EPI_ISL_689665\|2020-07 | .................... | .................... | .................... | .................... | .................... | .................... |
| V | 2987 | hCoV-19/Japan/PG-4528/2020\|EPI_ISL_689666\|2020-07 | .................... | .................... | .................... | .................... | .................... | .................... |
| V | 2988 | hCoV-19/Japan/PG-4529/2020\|EPI_ISL_689667\|2020-07 | .................... | .................... | .................... | .................... | .................... | .................... |
| V | 2989 | hCoV-19/Japan/PG-4530/2020\|EPI_ISL_689668\|2020-07 | .................... | .................... | .................... | .................... | .................... | .................... |
| V | 2990 | hCoV-19/Japan/PG-4531/2020\|EPI_ISL_689669\|2020-07 | .................... | .................... | .................... | .................... | .................... | .................... |
| V | 2991 | hCoV-19/Japan/PG-5498/2020\|EPI_ISL_690600\|2020-08 | .................... | .................... | .................... | .................... | .................... | .................... |
| V | 2992 | hCoV-19/Japan/PG-5499/2020\|EPI_ISL_690601\|2020-08 | .................... | .................... | .................... | .................... | .................... | .................... |
| V | 2993 | hCoV-19/Japan/PG-5500/2020\|EPI_ISL_690602\|2020-08 | .................... | .................... | .................... | .................... | .................... | .................... |
| V | 2994 | hCoV-19/Japan/PG-5501/2020\|EPI_ISL_690603\|2020-08 | .................... | .................... | .................... | .................... | .................... | .................... |
| V | 2995 | hCoV-19/Japan/PG-5502/2020\|EPI_ISL_690604\|2020-08 | .................... | .................... | .................... | .................... | .................... | .................... |
| V | 2996 | hCoV-19/Japan/PG-5503/2020\|EPI_ISL_690605\|2020-08 | .................... | .................... | .................... | .................... | .................... | .................... |
| V | 2997 | hCoV-19/Japan/PG-5504/2020\|EPI_ISL_690606\|2020-08 | .................... | .................... | .................... | .................... | .................... | .................... |
| V | 2998 | hCoV-19/Japan/PG-5505/2020\|EPI_ISL_690607\|2020-08 | .................... | .................... | .................... | .................... | .................... | .................... |
| V | 2999 | hCoV-19/Japan/PG-5506/2020\|EPI_ISL_690608\|2020-08 | .................... | .................... | .................... | .................... | .................... | .................... |
| V | 3000 | hCoV-19/Japan/PG-5507/2020\|EPI_ISL_690609\|2020-08 | .................... | .................... | .................... | .................... | .................... | .................... |
| V | 3001 | hCoV-19/Japan/PG-5508/2020\|EPI_ISL_690610\|2020-08 | .................... | .................... | .................... | .................... | .................... | .................... |
| V | 3002 | hCoV-19/Japan/PG-5509/2020\|EPI_ISL_690611\|2020-08 | .................... | .................... | .................... | .................... | .................... | .................... |
| V | 3003 | hCoV-19/Japan/PG-2969/2020\|EPI_ISL_690614\|2020-04 | .................... | .................... | .................... | .................... | .................... | .................... |
| V | 3004 | hCoV-19/Japan/PG-2978/2020\|EPI_ISL_690615\|2020-04 | .................... | .................... | .................... | .................... | .................... | .................... |
| V | 3005 | hCoV-19/Japan/PG-3438/2020\|EPI_ISL_690616\|2020-06 | .................... | .................... | .................... | .................... | .................... | .................... |
| V | 3006 | hCoV-19/Japan/PG-3087/2020\|EPI_ISL_690617\|2020-04 | .................... | .................... | .................... | .................... | .................... | .................... |
| V | 3007 | hCoV-19/Japan/PG-2520/2020\|EPI_ISL_690619\|2020-05 | .................... | .................... | .................... | .................... | .................... | .................... |
| V | 3008 | hCoV-19/Japan/PG-3205/2020\|EPI_ISL_690620\|2020-04 | .................... | .................... | .................... | .................... | .................... | .................... |
| V | 3009 | hCoV-19/Japan/PG-3207/2020\|EPI_ISL_690621\|2020-04 | .................... | .................... | .................... | .................... | .................... | .................... |
| V | 3010 | hCoV-19/Japan/PG-3223/2020\|EPI_ISL_690622\|2020-04 | .................... | .................... | .................... | .................... | .................... | .................... |
| V | 3011 | hCoV-19/Japan/PG-3245/2020\|EPI_ISL_690623\|2020-04 | .................... | .................... | .................... | .................... | .................... | .................... |
| V | 3012 | hCoV-19/Japan/PG-3387/2020\|EPI_ISL_690624\|2020-05 | .................... | .................... | .................... | .................... | .................... | .................... |
| V | 3013 | hCoV-19/Japan/PG-3233/2020\|EPI_ISL_690625\|2020-04 | .................... | .................... | .................... | .................... | .................... | .................... |
| V | 3014 | hCoV-19/Japan/PG-3149/2020\|EPI_ISL_690635\|2020-06 | .................... | .................... | .................... | .................... | .................... | .................... |
| V | 3015 | hCoV-19/Japan/PG-2552/2020\|EPI_ISL_690687\|2020-05 | .................... | .................... | .................... | .................... | .................... | .................... |
| V | 3016 | hCoV-19/Japan/PG-2553/2020\|EPI_ISL_690688\|2020-05 | .................... | .................... | .................... | .................... | .................... | .................... |
| V | 3017 | hCoV-19/Japan/PG-2555/2020\|EPI_ISL_690690\|2020-06 | .................... | .................... | .................... | .................... | .................... | .................... |

| | | | | | | | | |
|---|---|---|---|---|---|---|---|---|
| V | 3018 | hCoV-19/Japan/PG-2556/2020\|EPI_ISL_690691\|2020-06 | .................... | .................... | .................... | .................... | .................... | .................... |
| V | 3019 | hCoV-19/Japan/PG-2557/2020\|EPI_ISL_690692\|2020-06 | .................... | .................... | .................... | .................... | .................... | .................... |
| V | 3020 | hCoV-19/Japan/PG-2558/2020\|EPI_ISL_690693\|2020-06 | .................... | .................... | .................... | .................... | .................... | .................... |
| V | 3021 | hCoV-19/Japan/PG-2559/2020\|EPI_ISL_690694\|2020-06 | .................... | .................... | .................... | .................... | .................... | .................... |
| V | 3022 | hCoV-19/Japan/PG-2560/2020\|EPI_ISL_690695\|2020-06 | .................... | .................... | .................... | .................... | .................... | .................... |
| V | 3023 | hCoV-19/Japan/PG-2561/2020\|EPI_ISL_690696\|2020-06 | .................... | .................... | .................... | .................... | .................... | .................... |
| V | 3024 | hCoV-19/Japan/PG-2562/2020\|EPI_ISL_690697\|2020-06 | .................... | .................... | .................... | .................... | .................... | .................... |
| V | 3025 | hCoV-19/Japan/PG-2563/2020\|EPI_ISL_690698\|2020-06 | .................... | .................... | .................... | .................... | .................... | .................... |
| V | 3026 | hCoV-19/Japan/PG-2564/2020\|EPI_ISL_690699\|2020-06 | .................... | .................... | .................... | .................... | .................... | .................... |
| V | 3027 | hCoV-19/Japan/PG-3667/2020\|EPI_ISL_691920\|2020-04 | .................... | .................... | .................... | .................... | .................... | .................... |
| V | 3028 | hCoV-19/Japan/PG-3668/2020\|EPI_ISL_691921\|2020-04 | .................... | .................... | .................... | .................... | .................... | .................... |
| V | 3029 | hCoV-19/Japan/PG-3669/2020\|EPI_ISL_691922\|2020-04 | .................... | .................... | .................... | .................... | .................... | .................... |
| V | 3030 | hCoV-19/Japan/PG-3670/2020\|EPI_ISL_691923\|2020-04 | .................... | .................... | .................... | .................... | .................... | .................... |
| V | 3031 | hCoV-19/Japan/PG-3671/2020\|EPI_ISL_691924\|2020-04 | .................... | .................... | .................... | .................... | .................... | .................... |
| V | 3032 | hCoV-19/Japan/PG-3672/2020\|EPI_ISL_691925\|2020-04 | .................... | .................... | .................... | .................... | .................... | .................... |
| V | 3033 | hCoV-19/Japan/PG-3673/2020\|EPI_ISL_691926\|2020-04 | .................... | .................... | .................... | .................... | .................... | .................... |
| V | 3034 | hCoV-19/Japan/PG-3674/2020\|EPI_ISL_691927\|2020-04 | .................... | .................... | .................... | .................... | .................... | .................... |
| V | 3035 | hCoV-19/Japan/PG-3675/2020\|EPI_ISL_691928\|2020-04 | .................... | .................... | .................... | .................... | .................... | .................... |
| V | 3036 | hCoV-19/Japan/PG-3676/2020\|EPI_ISL_691929\|2020-04 | .................... | .................... | .................... | .................... | .................... | .................... |
| V | 3037 | hCoV-19/Japan/PG-3677/2020\|EPI_ISL_691930\|2020-04 | .................... | .................... | .................... | .................... | .................... | .................... |
| V | 3038 | hCoV-19/Japan/PG-3678/2020\|EPI_ISL_691931\|2020-04 | .................... | .................... | .................... | .................... | .................... | .................... |
| V | 3039 | hCoV-19/Japan/PG-3679/2020\|EPI_ISL_691932\|2020-04 | .................... | .................... | .................... | .................... | .................... | .................... |
| V | 3040 | hCoV-19/Japan/PG-3680/2020\|EPI_ISL_691933\|2020-04 | .................... | .................... | .................... | .................... | .................... | .................... |
| V | 3041 | hCoV-19/Japan/PG-3681/2020\|EPI_ISL_691934\|2020-04 | .................... | .................... | .................... | .................... | .................... | .................... |
| V | 3042 | hCoV-19/Japan/PG-3682/2020\|EPI_ISL_691935\|2020-04 | .................... | .................... | .................... | .................... | .................... | .................... |
| V | 3043 | hCoV-19/Japan/PG-3683/2020\|EPI_ISL_691936\|2020-04 | .................... | .................... | .................... | .................... | .................... | .................... |
| V | 3044 | hCoV-19/Japan/PG-3684/2020\|EPI_ISL_691937\|2020-04 | .................... | .................... | .................... | .................... | .................... | .................... |
| V | 3045 | hCoV-19/Japan/PG-3685/2020\|EPI_ISL_691938\|2020-04 | .................... | .................... | .................... | .................... | .................... | .................... |
| V | 3046 | hCoV-19/Japan/PG-3686/2020\|EPI_ISL_691939\|2020-04 | .................... | .................... | .................... | .................... | .................... | .................... |
| V | 3047 | hCoV-19/Japan/PG-3687/2020\|EPI_ISL_691940\|2020-04 | .................... | .................... | .................... | .................... | .................... | .................... |
| V | 3048 | hCoV-19/Japan/PG-3688/2020\|EPI_ISL_691941\|2020-04 | .................... | .................... | .................... | .................... | .................... | .................... |
| V | 3049 | hCoV-19/Japan/PG-3689/2020\|EPI_ISL_691942\|2020-04 | .................... | .................... | .................... | .................... | .................... | .................... |
| V | 3050 | hCoV-19/Japan/PG-3690/2020\|EPI_ISL_691943\|2020-04 | .................... | .................... | .................... | .................... | .................... | .................... |
| V | 3051 | hCoV-19/Japan/PG-3691/2020\|EPI_ISL_691944\|2020-04 | .................... | .................... | .................... | .................... | .................... | .................... |
| V | 3052 | hCoV-19/Japan/PG-3692/2020\|EPI_ISL_691945\|2020-04 | .................... | .................... | .................... | .................... | .................... | .................... |
| V | 3053 | hCoV-19/Japan/PG-3693/2020\|EPI_ISL_691946\|2020-04 | .................... | .................... | .................... | .................... | .................... | .................... |
| V | 3054 | hCoV-19/Japan/PG-3694/2020\|EPI_ISL_691947\|2020-04 | .................... | .................... | .................... | .................... | .................... | .................... |
| V | 3055 | hCoV-19/Japan/PG-3695/2020\|EPI_ISL_691948\|2020-04 | .................... | .................... | .................... | .................... | .................... | .................... |
| V | 3056 | hCoV-19/Japan/PG-3696/2020\|EPI_ISL_691949\|2020-04 | .................... | .................... | .................... | .................... | .................... | .................... |
| V | 3057 | hCoV-19/Japan/PG-3697/2020\|EPI_ISL_691950\|2020-04 | .................... | .................... | .................... | .................... | .................... | .................... |
| V | 3058 | hCoV-19/Japan/PG-3698/2020\|EPI_ISL_691951\|2020-04 | .................... | .................... | .................... | .................... | .................... | .................... |
| V | 3059 | hCoV-19/Japan/PG-3699/2020\|EPI_ISL_691952\|2020-04 | .................... | .................... | .................... | .................... | .................... | .................... |
| V | 3060 | hCoV-19/Japan/PG-3700/2020\|EPI_ISL_691953\|2020-04 | .................... | .................... | .................... | .................... | .................... | .................... |
| V | 3061 | hCoV-19/Japan/PG-3701/2020\|EPI_ISL_691954\|2020-04 | .................... | .................... | .................... | .................... | .................... | .................... |
| V | 3062 | hCoV-19/Japan/PG-3702/2020\|EPI_ISL_691955\|2020-04 | .................... | .................... | .................... | .................... | .................... | .................... |
| V | 3063 | hCoV-19/Japan/PG-3703/2020\|EPI_ISL_691956\|2020-04 | .................... | .................... | .................... | .................... | .................... | .................... |
| V | 3064 | hCoV-19/Japan/PG-3704/2020\|EPI_ISL_691957\|2020-04 | .................... | .................... | .................... | .................... | .................... | .................... |
| V | 3065 | hCoV-19/Japan/PG-3705/2020\|EPI_ISL_691958\|2020-04 | .................... | .................... | .................... | .................... | .................... | .................... |
| V | 3066 | hCoV-19/Japan/PG-3706/2020\|EPI_ISL_691959\|2020-04 | .................... | .................... | .................... | .................... | .................... | .................... |
| V | 3067 | hCoV-19/Japan/PG-3707/2020\|EPI_ISL_691960\|2020-04 | .................... | .................... | .................... | .................... | .................... | .................... |
| V | 3068 | hCoV-19/Japan/PG-3712/2020\|EPI_ISL_691965\|2020-04 | .................... | .................... | .................... | .................... | .................... | .................... |
| V | 3069 | hCoV-19/Japan/PG-3713/2020\|EPI_ISL_691966\|2020-04 | .................... | .................... | .................... | .................... | .................... | .................... |
| V | 3070 | hCoV-19/Japan/PG-3714/2020\|EPI_ISL_691967\|2020-04 | .................... | .................... | .................... | .................... | .................... | .................... |
| V | 3071 | hCoV-19/Japan/PG-3715/2020\|EPI_ISL_691968\|2020-04 | .................... | .................... | .................... | .................... | .................... | .................... |
| V | 3072 | hCoV-19/Japan/PG-3716/2020\|EPI_ISL_691969\|2020-04 | .................... | .................... | .................... | .................... | .................... | .................... |
| V | 3073 | hCoV-19/Japan/PG-1100/2020\|EPI_ISL_684710\|2020-04 | .................... | .................... | .................... | .................... | .................... | .................... |
| V | 3074 | hCoV-19/Japan/PG-1101/2020\|EPI_ISL_684711\|2020-04 | .................... | .................... | .................... | .................... | .................... | .................... |
| V | 3075 | hCoV-19/Japan/PG-1102/2020\|EPI_ISL_684712\|2020-04 | .................... | .................... | .................... | .................... | .................... | .................... |
| V | 3076 | hCoV-19/Japan/PG-1103/2020\|EPI_ISL_684713\|2020-04 | .................... | .................... | .................... | .................... | .................... | .................... |
| V | 3077 | hCoV-19/Japan/PG-1104/2020\|EPI_ISL_684714\|2020-04 | .................... | .................... | .................... | .................... | .................... | .................... |
| V | 3078 | hCoV-19/Japan/PG-1105/2020\|EPI_ISL_684715\|2020-04 | .................... | .................... | .................... | .................... | .................... | .................... |
| V | 3079 | hCoV-19/Japan/PG-1106/2020\|EPI_ISL_684716\|2020-03 | .................... | .................... | .................... | .................... | .................... | .................... |
| V | 3080 | hCoV-19/Japan/PG-1107/2020\|EPI_ISL_684717\|2020-03 | .................... | .................... | .................... | .................... | .................... | .................... |
| V | 3081 | hCoV-19/Japan/PG-1113/2020\|EPI_ISL_684723\|2020-04 | .................... | .................... | .................... | .................... | .................... | .................... |
| V | 3082 | hCoV-19/Japan/PG-1114/2020\|EPI_ISL_684724\|2020-04 | .................... | .................... | .................... | .................... | .................... | .................... |
| V | 3083 | hCoV-19/Japan/PG-1115/2020\|EPI_ISL_684725\|2020-04 | .................... | .................... | .................... | .................... | .................... | .................... |
| V | 3084 | hCoV-19/Japan/PG-1116/2020\|EPI_ISL_684726\|2020-04 | .................... | .................... | .................... | .................... | .................... | .................... |
| V | 3085 | hCoV-19/Japan/PG-1117/2020\|EPI_ISL_684727\|2020-04 | .................... | .................... | .................... | .................... | .................... | .................... |
| V | 3086 | hCoV-19/Japan/PG-1118/2020\|EPI_ISL_684728\|2020-04 | .................... | .................... | .................... | .................... | .................... | .................... |
| V | 3087 | hCoV-19/Japan/PG-1119/2020\|EPI_ISL_684729\|2020-04 | .................... | .................... | .................... | .................... | .................... | .................... |
| V | 3088 | hCoV-19/Japan/PG-1120/2020\|EPI_ISL_684730\|2020-04 | .................... | .................... | .................... | .................... | .................... | .................... |
| V | 3089 | hCoV-19/Japan/PG-1121/2020\|EPI_ISL_684731\|2020-04 | .................... | .................... | .................... | .................... | .................... | .................... |
| V | 3090 | hCoV-19/Japan/PG-1122/2020\|EPI_ISL_684732\|2020-04 | .................... | .................... | .................... | .................... | .................... | .................... |
| V | 3091 | hCoV-19/Japan/PG-1124/2020\|EPI_ISL_684734\|2020-04 | .................... | .................... | .................... | .................... | .................... | .................... |
| V | 3092 | hCoV-19/Japan/PG-1125/2020\|EPI_ISL_684735\|2020-04 | .................... | .................... | .................... | .................... | .................... | .................... |
| V | 3093 | hCoV-19/Japan/PG-1126/2020\|EPI_ISL_684736\|2020-03 | .................... | .................... | .................... | .................... | .................... | .................... |
| V | 3094 | hCoV-19/Japan/PG-1127/2020\|EPI_ISL_684737\|2020-04 | .................... | .................... | .................... | .................... | .................... | .................... |
| V | 3095 | hCoV-19/Japan/PG-1128/2020\|EPI_ISL_684738\|2020-04 | .................... | .................... | .................... | .................... | .................... | .................... |
| V | 3096 | hCoV-19/Japan/PG-1129/2020\|EPI_ISL_684739\|2020-04 | .................... | .................... | .................... | .................... | .................... | .................... |
| V | 3097 | hCoV-19/Japan/PG-1130/2020\|EPI_ISL_684740\|2020-04 | .................... | .................... | .................... | .................... | .................... | .................... |

| | | | | | | | | |
|---|---|---|---|---|---|---|---|---|
| V | 3098 | hCoV-19/Japan/PG-1131/2020\|EPI_ISL_684741\|2020-04 | ···················· | ···················· | ····················· | ···················· | ···················· | ···················· |
| V | 3099 | hCoV-19/Japan/PG-1132/2020\|EPI_ISL_684742\|2020-04 | ···················· | ···················· | ····················· | ···················· | ···················· | ···················· |
| V | 3100 | hCoV-19/Japan/PG-1133/2020\|EPI_ISL_684743\|2020-04 | ···················· | ···················· | ····················· | ···················· | ···················· | ···················· |
| V | 3101 | hCoV-19/Japan/PG-1134/2020\|EPI_ISL_684744\|2020-04 | ···················· | ···················· | ····················· | ···················· | ···················· | ···················· |
| V | 3102 | hCoV-19/Japan/PG-1135/2020\|EPI_ISL_684745\|2020-04 | ···················· | ···················· | ····················· | ···················· | ···················· | ···················· |
| V | 3103 | hCoV-19/Japan/PG-1136/2020\|EPI_ISL_684746\|2020-04 | ···················· | ···················· | ····················· | ···················· | ···················· | ···················· |
| V | 3104 | hCoV-19/Japan/PG-1137/2020\|EPI_ISL_684747\|2020-04 | ···················· | ···················· | ····················· | ···················· | ···················· | ···················· |
| V | 3105 | hCoV-19/Japan/PG-1138/2020\|EPI_ISL_684748\|2020-04 | ···················· | ···················· | ····················· | ···················· | ···················· | ···················· |
| V | 3106 | hCoV-19/Japan/PG-1139/2020\|EPI_ISL_684749\|2020-04 | ···················· | ···················· | ····················· | ···················· | ···················· | ···················· |
| V | 3107 | hCoV-19/Japan/PG-1140/2020\|EPI_ISL_684750\|2020-04 | ···················· | ···················· | ····················· | ···················· | ···················· | ···················· |
| V | 3108 | hCoV-19/Japan/PG-1141/2020\|EPI_ISL_684751\|2020-04 | ···················· | ···················· | ····················· | ···················· | ···················· | ···················· |
| V | 3109 | hCoV-19/Japan/PG-1142/2020\|EPI_ISL_684752\|2020-04 | ···················· | ···················· | ····················· | ···················· | ···················· | ···················· |
| V | 3110 | hCoV-19/Japan/PG-1143/2020\|EPI_ISL_684753\|2020-04 | ···················· | ···················· | ····················· | ···················· | ···················· | ···················· |
| V | 3111 | hCoV-19/Japan/PG-1144/2020\|EPI_ISL_684754\|2020-04 | ···················· | ···················· | ····················· | ···················· | ···················· | ···················· |
| V | 3112 | hCoV-19/Japan/PG-1145/2020\|EPI_ISL_684755\|2020-04 | ···················· | ···················· | ····················· | ···················· | ···················· | ···················· |
| V | 3113 | hCoV-19/Japan/PG-1146/2020\|EPI_ISL_684756\|2020-04 | ···················· | ···················· | ····················· | ···················· | ···················· | ···················· |
| V | 3114 | hCoV-19/Japan/PG-1147/2020\|EPI_ISL_684757\|2020-04 | ···················· | ···················· | ····················· | ···················· | ···················· | ···················· |
| V | 3115 | hCoV-19/Japan/PG-1148/2020\|EPI_ISL_684758\|2020-04 | ···················· | ···················· | ····················· | ···················· | ···················· | ···················· |
| V | 3116 | hCoV-19/Japan/PG-1149/2020\|EPI_ISL_684759\|2020-04 | ···················· | ···················· | ····················· | ···················· | ···················· | ···················· |
| V | 3117 | hCoV-19/Japan/PG-1150/2020\|EPI_ISL_684760\|2020-04 | ···················· | ···················· | ····················· | ···················· | ···················· | ···················· |
| V | 3118 | hCoV-19/Japan/PG-1151/2020\|EPI_ISL_684761\|2020-04 | ···················· | ···················· | ····················· | ···················· | ···················· | ···················· |
| V | 3119 | hCoV-19/Japan/PG-1152/2020\|EPI_ISL_684762\|2020-03 | ···················· | ···················· | ····················· | ···················· | ···················· | ···················· |
| V | 3120 | hCoV-19/Japan/PG-1153/2020\|EPI_ISL_684763\|2020-03 | ···················· | ···················· | ····················· | ···················· | ···················· | ···················· |
| V | 3121 | hCoV-19/Japan/PG-1154/2020\|EPI_ISL_684764\|2020-04 | ···················· | ···················· | ····················· | ···················· | ···················· | ···················· |
| V | 3122 | hCoV-19/Japan/PG-1155/2020\|EPI_ISL_684765\|2020-04 | ···················· | ···················· | ····················· | ···················· | ···················· | ···················· |
| V | 3123 | hCoV-19/Japan/PG-1156/2020\|EPI_ISL_684766\|2020-04 | ···················· | ···················· | ····················· | ···················· | ···················· | ···················· |
| V | 3124 | hCoV-19/Japan/PG-1157/2020\|EPI_ISL_684767\|2020-04 | ···················· | ···················· | ····················· | ···················· | ···················· | ···················· |
| V | 3125 | hCoV-19/Japan/PG-1158/2020\|EPI_ISL_684768\|2020-04 | ···················· | ···················· | ····················· | ···················· | ···················· | ···················· |
| V | 3126 | hCoV-19/Japan/PG-1159/2020\|EPI_ISL_684769\|2020-04 | ···················· | ···················· | ····················· | ···················· | ···················· | ···················· |
| V | 3127 | hCoV-19/Japan/PG-1160/2020\|EPI_ISL_684770\|2020-04 | ···················· | ···················· | ····················· | ···················· | ···················· | ···················· |
| V | 3128 | hCoV-19/Japan/PG-1161/2020\|EPI_ISL_684771\|2020-04 | ···················· | ···················· | ····················· | ···················· | ···················· | ···················· |
| V | 3129 | hCoV-19/Japan/PG-1162/2020\|EPI_ISL_684772\|2020-04 | ···················· | ···················· | ····················· | ···················· | ···················· | ···················· |
| V | 3130 | hCoV-19/Japan/PG-1163/2020\|EPI_ISL_684773\|2020-04 | ···················· | ···················· | ····················· | ···················· | ···················· | ···················· |
| V | 3131 | hCoV-19/Japan/PG-1164/2020\|EPI_ISL_684774\|2020-04 | ···················· | ···················· | ····················· | ···················· | ···················· | ···················· |
| V | 3132 | hCoV-19/Japan/PG-1165/2020\|EPI_ISL_684775\|2020-04 | ···················· | ···················· | ····················· | ···················· | ···················· | ···················· |
| V | 3133 | hCoV-19/Japan/PG-1166/2020\|EPI_ISL_684776\|2020-04 | ···················· | ···················· | ····················· | ···················· | ···················· | ···················· |
| V | 3134 | hCoV-19/Japan/PG-1167/2020\|EPI_ISL_684777\|2020-04 | ···················· | ···················· | ····················· | ···················· | ···················· | ···················· |
| V | 3135 | hCoV-19/Japan/PG-1168/2020\|EPI_ISL_684778\|2020-04 | ···················· | ···················· | ····················· | ···················· | ···················· | ···················· |
| V | 3136 | hCoV-19/Japan/PG-1169/2020\|EPI_ISL_684779\|2020-04 | ···················· | ···················· | ····················· | ···················· | ···················· | ···················· |
| V | 3137 | hCoV-19/Japan/PG-1170/2020\|EPI_ISL_684780\|2020-04 | ···················· | ···················· | ····················· | ···················· | ···················· | ···················· |
| V | 3138 | hCoV-19/Japan/PG-1171/2020\|EPI_ISL_684781\|2020-04 | ···················· | ···················· | ····················· | ···················· | ···················· | ···················· |
| V | 3139 | hCoV-19/Japan/PG-1172/2020\|EPI_ISL_684782\|2020-04 | ···················· | ···················· | ····················· | ···················· | ···················· | ···················· |
| V | 3140 | hCoV-19/Japan/PG-1173/2020\|EPI_ISL_684783\|2020-04 | ···················· | ···················· | ····················· | ···················· | ···················· | ···················· |
| V | 3141 | hCoV-19/Japan/PG-1174/2020\|EPI_ISL_684784\|2020-04 | ···················· | ···················· | ····················· | ···················· | ···················· | ···················· |
| V | 3142 | hCoV-19/Japan/PG-1175/2020\|EPI_ISL_684785\|2020-04 | ···················· | ···················· | ····················· | ···················· | ···················· | ···················· |
| V | 3143 | hCoV-19/Japan/PG-1176/2020\|EPI_ISL_684786\|2020-04 | ···················· | ···················· | ····················· | ···················· | ···················· | ···················· |
| V | 3144 | hCoV-19/Japan/PG-1177/2020\|EPI_ISL_684787\|2020-04 | ···················· | ···················· | ····················· | ···················· | ···················· | ···················· |
| V | 3145 | hCoV-19/Japan/PG-1178/2020\|EPI_ISL_684788\|2020-04 | ···················· | ···················· | ····················· | ···················· | ···················· | ···················· |
| V | 3146 | hCoV-19/Japan/PG-1179/2020\|EPI_ISL_684789\|2020-04 | ···················· | ···················· | ····················· | ···················· | ···················· | ···················· |
| V | 3147 | hCoV-19/Japan/PG-1180/2020\|EPI_ISL_684790\|2020-04 | ···················· | ····A··············· | ····················· | ···················· | ···················· | ···················· |
| V | 3148 | hCoV-19/Japan/PG-1181/2020\|EPI_ISL_684791\|2020-04 | ···················· | ···················· | ····················· | ···················· | ···················· | ···················· |
| V | 3149 | hCoV-19/Japan/PG-1182/2020\|EPI_ISL_684792\|2020-04 | ···················· | ···················· | ····················· | ···················· | ···················· | ···················· |
| V | 3150 | hCoV-19/Japan/PG-1183/2020\|EPI_ISL_684793\|2020-04 | ···················· | ···················· | ····················· | ···················· | ···················· | ···················· |
| V | 3151 | hCoV-19/Japan/PG-1184/2020\|EPI_ISL_684794\|2020-04 | ···················· | ···················· | ····················· | ···················· | ···················· | ···················· |
| V | 3152 | hCoV-19/Japan/PG-1185/2020\|EPI_ISL_684795\|2020-04 | ···················· | ···················· | ····················· | ···················· | ···················· | ···················· |
| V | 3153 | hCoV-19/Japan/PG-1186/2020\|EPI_ISL_684796\|2020-04 | ···················· | ···················· | ····················· | ···················· | ···················· | ···················· |
| V | 3154 | hCoV-19/Japan/PG-1187/2020\|EPI_ISL_684797\|2020-04 | ···················· | ···················· | ····················· | ···················· | ···················· | ···················· |
| V | 3155 | hCoV-19/Japan/PG-1188/2020\|EPI_ISL_684798\|2020-03 | ···················· | ···················· | ····················· | ···················· | ···················· | ···················· |
| V | 3156 | hCoV-19/Japan/PG-1189/2020\|EPI_ISL_684799\|2020-03 | ···················· | ···················· | ····················· | ···················· | ···················· | ···················· |
| V | 3157 | hCoV-19/Japan/PG-7928/2020\|EPI_ISL_687029\|2020-07 | ···················· | ···················· | ····················· | ···················· | ···················· | ···················· |
| V | 3158 | hCoV-19/Japan/PG-7929/2020\|EPI_ISL_687030\|2020-07 | ···················· | ···················· | ····················· | ···················· | ···················· | ···················· |
| V | 3159 | hCoV-19/Japan/PG-7930/2020\|EPI_ISL_687031\|2020-07 | ···················· | ···················· | ····················· | ···················· | ···················· | ···················· |
| V | 3160 | hCoV-19/Japan/PG-7931/2020\|EPI_ISL_687032\|2020-07 | ···················· | ···················· | ····················· | ···················· | ···················· | ···················· |
| V | 3161 | hCoV-19/Japan/PG-7932/2020\|EPI_ISL_687033\|2020-07 | ···················· | ···················· | ····················· | ···················· | ···················· | ···················· |
| V | 3162 | hCoV-19/Japan/PG-7933/2020\|EPI_ISL_687034\|2020-07 | ···················· | ···················· | ····················· | ···················· | ···················· | ···················· |
| V | 3163 | hCoV-19/Japan/PG-7934/2020\|EPI_ISL_687035\|2020-07 | ···················· | ···················· | ····················· | ···················· | ···················· | ···················· |
| V | 3164 | hCoV-19/Japan/PG-7935/2020\|EPI_ISL_687036\|2020-07 | ···················· | ···················· | ····················· | ···················· | ···················· | ···················· |
| V | 3165 | hCoV-19/Japan/PG-7936/2020\|EPI_ISL_687037\|2020-07 | ···················· | ···················· | ····················· | ···················· | ···················· | ···················· |
| V | 3166 | hCoV-19/Japan/PG-7937/2020\|EPI_ISL_687038\|2020-07 | ···················· | ···················· | ····················· | ···················· | ···················· | ···················· |
| V | 3167 | hCoV-19/Japan/PG-7938/2020\|EPI_ISL_687039\|2020-07 | ···················· | ···················· | ····················· | ···················· | ···················· | ···················· |
| V | 3168 | hCoV-19/Japan/PG-7939/2020\|EPI_ISL_687040\|2020-07 | ···················· | ···················· | ····················· | ···················· | ···················· | ···················· |
| V | 3169 | hCoV-19/Japan/PG-7940/2020\|EPI_ISL_687041\|2020-07 | ···················· | ···················· | ····················· | ···················· | ···················· | ···················· |
| V | 3170 | hCoV-19/Japan/PG-7941/2020\|EPI_ISL_687042\|2020-08 | ···················· | ···················· | ····················· | ···················· | ···················· | ···················· |
| V | 3171 | hCoV-19/Japan/PG-7942/2020\|EPI_ISL_687043\|2020-05 | ···················· | ···················· | ····················· | ···················· | ···················· | ···················· |
| V | 3172 | hCoV-19/Japan/PG-7943/2020\|EPI_ISL_687044\|2020-05 | ···················· | ···················· | ····················· | ···················· | ···················· | ···················· |
| V | 3173 | hCoV-19/Japan/PG-7944/2020\|EPI_ISL_687045\|2020-05 | ···················· | ···················· | ····················· | ···················· | ···················· | ···················· |
| V | 3174 | hCoV-19/Japan/PG-7945/2020\|EPI_ISL_687046\|2020-05 | ···················· | ···················· | ····················· | ···················· | ···················· | ···················· |
| V | 3175 | hCoV-19/Japan/PG-7946/2020\|EPI_ISL_687047\|2020-05 | ···················· | ···················· | ····················· | ···················· | ···················· | ···················· |
| V | 3176 | hCoV-19/Japan/PG-7947/2020\|EPI_ISL_687048\|2020-05 | ···················· | ···················· | ····················· | ···················· | ···················· | ···················· |
| V | 3177 | hCoV-19/Japan/PG-7948/2020\|EPI_ISL_687049\|2020-05 | ···················· | ···················· | ····················· | ···················· | ···················· | ···················· |

| | | | | | | | | |
|---|---|---|---|---|---|---|---|---|
| V | 3178 | hCoV-19/Japan/PG-7949/2020\|EPI_ISL_687050\|2020-05 | .................... | .................... | ..................... | .................... | .................... | .................... |
| V | 3179 | hCoV-19/Japan/PG-7950/2020\|EPI_ISL_687051\|2020-05 | .................... | .................... | ..................... | .................... | .................... | .................... |
| V | 3180 | hCoV-19/Japan/PG-7951/2020\|EPI_ISL_687052\|2020-05 | .................... | .................... | ..................... | .................... | .................... | .................... |
| V | 3181 | hCoV-19/Japan/PG-7952/2020\|EPI_ISL_687053\|2020-05 | .................... | .................... | ..................... | .................... | .................... | .................... |
| V | 3182 | hCoV-19/Japan/PG-7953/2020\|EPI_ISL_687054\|2020-05 | .................... | .................... | ..................... | .................... | .................... | .................... |
| V | 3183 | hCoV-19/Japan/PG-7954/2020\|EPI_ISL_687055\|2020-05 | .................... | .................... | ..................... | .................... | .................... | .................... |
| V | 3184 | hCoV-19/Japan/PG-7955/2020\|EPI_ISL_687056\|2020-05 | .................... | .................... | ..................... | .................... | .................... | .................... |
| V | 3185 | hCoV-19/Japan/PG-7956/2020\|EPI_ISL_687057\|2020-05 | .................... | .................... | ..................... | .................... | .................... | .................... |
| V | 3186 | hCoV-19/Japan/PG-7957/2020\|EPI_ISL_687058\|2020-05 | .................... | .................... | ..................... | .................... | .................... | .................... |
| V | 3187 | hCoV-19/Japan/PG-7958/2020\|EPI_ISL_687059\|2020-05 | .................... | .................... | ..................... | .................... | .................... | .................... |
| V | 3188 | hCoV-19/Japan/PG-7959/2020\|EPI_ISL_687060\|2020-05 | .................... | .................... | ..................... | .................... | .................... | .................... |
| V | 3189 | hCoV-19/Japan/PG-7960/2020\|EPI_ISL_687061\|2020-05 | .................... | .................... | ..................... | .................... | .................... | .................... |
| V | 3190 | hCoV-19/Japan/PG-7961/2020\|EPI_ISL_687062\|2020-05 | .................... | .................... | ..................... | .................... | .................... | .................... |
| V | 3191 | hCoV-19/Japan/PG-7132/2020\|EPI_ISL_688237\|2020-08 | .................... | .................... | ..................... | .................... | .................... | .................... |
| V | 3192 | hCoV-19/Japan/PG-7133/2020\|EPI_ISL_688238\|2020-08 | .................... | .................... | ..................... | .................... | .................... | .................... |
| V | 3193 | hCoV-19/Japan/PG-7257/2020\|EPI_ISL_688360\|2020-08 | .................... | .................... | ..................... | .................... | .................... | .................... |
| V | 3194 | hCoV-19/Japan/PG-7258/2020\|EPI_ISL_688361\|2020-08 | .................... | .................... | ..................... | .................... | .................... | .................... |
| V | 3195 | hCoV-19/Japan/PG-7259/2020\|EPI_ISL_688362\|2020-08 | .................... | .................... | ..................... | .................... | .................... | .................... |
| V | 3196 | hCoV-19/Japan/PG-7260/2020\|EPI_ISL_688363\|2020-08 | .................... | .................... | ..................... | .................... | .................... | .................... |
| V | 3197 | hCoV-19/Japan/PG-7261/2020\|EPI_ISL_688364\|2020-08 | .................... | .................... | ..................... | .................... | .................... | .................... |
| V | 3198 | hCoV-19/Japan/PG-7262/2020\|EPI_ISL_688365\|2020-08 | .................... | .................... | ..................... | .................... | .................... | .................... |
| V | 3199 | hCoV-19/Japan/PG-7263/2020\|EPI_ISL_688366\|2020-08 | .................... | .................... | ..................... | .................... | .................... | .................... |
| V | 3200 | hCoV-19/Japan/PG-7264/2020\|EPI_ISL_688367\|2020-08 | .................... | .................... | ..................... | .................... | .................... | .................... |
| V | 3201 | hCoV-19/Japan/PG-7265/2020\|EPI_ISL_688368\|2020-08 | .................... | .................... | ..................... | .................... | .................... | .................... |
| V | 3202 | hCoV-19/Japan/PG-7266/2020\|EPI_ISL_688369\|2020-08 | .................... | .................... | ..................... | .................... | .................... | .................... |
| V | 3203 | hCoV-19/Japan/PG-7267/2020\|EPI_ISL_688370\|2020-08 | .................... | .................... | ..................... | .................... | .................... | .................... |
| V | 3204 | hCoV-19/Japan/PG-7268/2020\|EPI_ISL_688371\|2020-08 | .................... | .................... | ..................... | .................... | .................... | .................... |
| V | 3205 | hCoV-19/Japan/PG-7269/2020\|EPI_ISL_688372\|2020-08 | .................... | .................... | ..................... | .................... | .................... | .................... |
| V | 3206 | hCoV-19/Japan/PG-7270/2020\|EPI_ISL_688373\|2020-08 | .................... | .................... | ..................... | .................... | .................... | .................... |
| V | 3207 | hCoV-19/Japan/PG-7271/2020\|EPI_ISL_688374\|2020-08 | .................... | .................... | ..................... | .................... | .................... | .................... |
| V | 3208 | hCoV-19/Japan/PG-7272/2020\|EPI_ISL_688375\|2020-08 | .................... | .................... | ..................... | .................... | .................... | .................... |
| V | 3209 | hCoV-19/Japan/PG-7273/2020\|EPI_ISL_688376\|2020-08 | .................... | .................... | ..................... | .................... | .................... | .................... |
| V | 3210 | hCoV-19/Japan/PG-7274/2020\|EPI_ISL_688377\|2020-08 | .................... | .................... | ..................... | .................... | .................... | .................... |
| V | 3211 | hCoV-19/Japan/PG-7275/2020\|EPI_ISL_688378\|2020-08 | .................... | .................... | ..................... | .................... | .................... | .................... |
| V | 3212 | hCoV-19/Japan/PG-7276/2020\|EPI_ISL_688379\|2020-08 | .................... | .................... | ..................... | .................... | .................... | .................... |
| V | 3213 | hCoV-19/Japan/PG-7277/2020\|EPI_ISL_688380\|2020-08 | .................... | .................... | ..................... | .................... | .................... | .................... |
| V | 3214 | hCoV-19/Japan/PG-7278/2020\|EPI_ISL_688381\|2020-08 | .................... | .................... | ..................... | .................... | .................... | .................... |
| V | 3215 | hCoV-19/Japan/PG-7279/2020\|EPI_ISL_688382\|2020-08 | .................... | .................... | ..................... | .................... | .................... | .................... |
| V | 3216 | hCoV-19/Japan/PG-7280/2020\|EPI_ISL_688383\|2020-08 | .................... | .................... | ..................... | .................... | .................... | .................... |
| V | 3217 | hCoV-19/Japan/PG-7281/2020\|EPI_ISL_688384\|2020-08 | .................... | .................... | ..................... | .................... | .................... | .................... |
| V | 3218 | hCoV-19/Japan/PG-7282/2020\|EPI_ISL_688385\|2020-08 | .................... | .................... | ..................... | .................... | .................... | .................... |
| V | 3219 | hCoV-19/Japan/PG-7283/2020\|EPI_ISL_688386\|2020-08 | .................... | .................... | ..................... | .................... | .................... | .................... |
| V | 3220 | hCoV-19/Japan/PG-7284/2020\|EPI_ISL_688387\|2020-08 | .................... | .................... | ..................... | .................... | .................... | .................... |
| V | 3221 | hCoV-19/Japan/PG-7285/2020\|EPI_ISL_688388\|2020-08 | .................... | .................... | ..................... | .................... | .................... | .................... |
| V | 3222 | hCoV-19/Japan/PG-7286/2020\|EPI_ISL_688389\|2020-08 | .................... | .................... | ..................... | .................... | .................... | .................... |
| V | 3223 | hCoV-19/Japan/PG-7287/2020\|EPI_ISL_688390\|2020-08 | .................... | .................... | ..................... | .................... | .................... | .................... |
| V | 3224 | hCoV-19/Japan/PG-7288/2020\|EPI_ISL_688391\|2020-08 | .................... | .................... | ..................... | .................... | .................... | .................... |
| V | 3225 | hCoV-19/Japan/PG-7289/2020\|EPI_ISL_688392\|2020-08 | .................... | .................... | ..................... | .................... | .................... | .................... |
| V | 3226 | hCoV-19/Japan/PG-6463/2020\|EPI_ISL_689568\|2020-08 | .................... | .................... | ..................... | .................... | .................... | .................... |
| V | 3227 | hCoV-19/Japan/PG-6464/2020\|EPI_ISL_689569\|2020-08 | .................... | .................... | ..................... | .................... | .................... | .................... |
| V | 3228 | hCoV-19/Japan/PG-6498/2020\|EPI_ISL_689600\|2020-09 | .................... | .................... | ..................... | .................... | .................... | .................... |
| V | 3229 | hCoV-19/Japan/PG-6499/2020\|EPI_ISL_689601\|2020-07 | .................... | .................... | ..................... | .................... | .................... | .................... |
| V | 3230 | hCoV-19/Japan/PG-6500/2020\|EPI_ISL_689602\|2020-07 | .................... | .................... | ..................... | .................... | .................... | .................... |
| V | 3232 | hCoV-19/Japan/PG-4553/2020\|EPI_ISL_689691\|2020 | .................... | .................... | ..................... | .................... | .................... | .................... |
| V | 3233 | hCoV-19/Japan/PG-4554/2020\|EPI_ISL_689692\|2020-07 | .................... | .................... | ..................... | .................... | .................... | .................... |
| V | 3234 | hCoV-19/Japan/PG-4555/2020\|EPI_ISL_689693\|2020-07 | .................... | .................... | ..................... | .................... | .................... | .................... |
| V | 3235 | hCoV-19/Japan/PG-4556/2020\|EPI_ISL_689694\|2020-07 | .................... | .................... | ..................... | .................... | .................... | .................... |
| V | 3236 | hCoV-19/Japan/PG-4558/2020\|EPI_ISL_689695\|2020-07 | .................... | .................... | ..................... | .................... | .................... | .................... |
| V | 3237 | hCoV-19/Japan/PG-4559/2020\|EPI_ISL_689696\|2020-07 | .................... | .................... | ..................... | .................... | .................... | .................... |
| V | 3238 | hCoV-19/Japan/PG-4560/2020\|EPI_ISL_689697\|2020-07 | .................... | .................... | ..................... | .................... | .................... | .................... |
| V | 3239 | hCoV-19/Japan/PG-4561/2020\|EPI_ISL_689698\|2020-07 | .................... | .................... | ..................... | .................... | .................... | .................... |
| V | 3240 | hCoV-19/Japan/PG-4562/2020\|EPI_ISL_689699\|2020-07 | .................... | .................... | ..................... | .................... | .................... | .................... |
| V | 3241 | hCoV-19/Japan/PG-5454/2020\|EPI_ISL_690557\|2020-08 | .................... | .................... | ..................... | .................... | .................... | .................... |
| V | 3242 | hCoV-19/Japan/PG-5455/2020\|EPI_ISL_690558\|2020-08 | .................... | .................... | ..................... | .................... | .................... | .................... |
| V | 3243 | hCoV-19/Japan/PG-2510/2020\|EPI_ISL_690631\|2020-04 | .................... | .................... | ..................... | .................... | .................... | .................... |
| V | 3244 | hCoV-19/Japan/PG-3146/2020\|EPI_ISL_690632\|2020-06 | .................... | .................... | ..................... | .................... | .................... | .................... |
| V | 3245 | hCoV-19/Japan/PG-3489/2020\|EPI_ISL_690633\|2020-07 | .................... | .................... | ..................... | .................... | .................... | .................... |
| V | 3246 | hCoV-19/Japan/PG-2503/2020\|EPI_ISL_690641\|2020-04 | .................... | .................... | ..................... | .................... | .................... | .................... |
| V | 3247 | hCoV-19/Japan/PG-2504/2020\|EPI_ISL_690642\|2020-04 | .................... | .................... | ..................... | .................... | .................... | .................... |
| V | 3248 | hCoV-19/Japan/PG-2505/2020\|EPI_ISL_690643\|2020-04 | .................... | .................... | ..................... | .................... | .................... | .................... |
| V | 3249 | hCoV-19/Japan/PG-2506/2020\|EPI_ISL_690644\|2020-04 | .................... | .................... | ..................... | .................... | .................... | .................... |
| V | 3250 | hCoV-19/Japan/PG-2507/2020\|EPI_ISL_690645\|2020-04 | .................... | .................... | ..................... | .................... | .................... | .................... |
| V | 3251 | hCoV-19/Japan/PG-2508/2020\|EPI_ISL_690646\|2020-04 | .................... | .................... | ..................... | .................... | .................... | .................... |
| V | 3252 | hCoV-19/Japan/PG-2509/2020\|EPI_ISL_690647\|2020-04 | .................... | .................... | ..................... | .................... | .................... | .................... |
| V | 3253 | hCoV-19/Japan/PG-2511/2020\|EPI_ISL_690648\|2020-04 | .................... | .................... | ..................... | .................... | .................... | .................... |
| V | 3254 | hCoV-19/Japan/PG-2512/2020\|EPI_ISL_690649\|2020-04 | .................... | .................... | ..................... | .................... | .................... | .................... |
| V | 3255 | hCoV-19/Japan/PG-2513/2020\|EPI_ISL_690650\|2020-04 | .................... | .................... | ..................... | .................... | .................... | .................... |
| V | 3256 | hCoV-19/Japan/PG-2515/2020\|EPI_ISL_690651\|2020-04 | .................... | .................... | ..................... | .................... | .................... | .................... |
| V | 3257 | hCoV-19/Japan/PG-2516/2020\|EPI_ISL_690652\|2020-04 | .................... | .................... | ..................... | .................... | .................... | .................... |
| V | 3258 | hCoV-19/Japan/PG-2517/2020\|EPI_ISL_690653\|2020-04 | .................... | .................... | ..................... | .................... | .................... | .................... |

| | | | | | | | | |
|---|---|---|---|---|---|---|---|---|
| V | 3259 | hCoV-19/Japan/PG-2518/2020\|EPI_ISL_690654\|2020-04 | .................... | .................... | .................... | .................... | .................... | .................... |
| V | 3260 | hCoV-19/Japan/PG-2519/2020\|EPI_ISL_690655\|2020-04 | .................... | .................... | .................... | .................... | .................... | .................... |
| V | 3261 | hCoV-19/Japan/PG-2521/2020\|EPI_ISL_690656\|2020-05 | .................... | .................... | .................... | .................... | .................... | .................... |
| V | 3262 | hCoV-19/Japan/PG-2522/2020\|EPI_ISL_690657\|2020-04 | .................... | .................... | .................... | .................... | .................... | .................... |
| V | 3263 | hCoV-19/Japan/PG-2523/2020\|EPI_ISL_690658\|2020-05 | .................... | .................... | .................... | .................... | .................... | .................... |
| V | 3264 | hCoV-19/Japan/PG-2524/2020\|EPI_ISL_690659\|2020-05 | .................... | .................... | .................... | .................... | .................... | .................... |
| V | 3265 | hCoV-19/Japan/PG-2525/2020\|EPI_ISL_690660\|2020-05 | .................... | .................... | .................... | .................... | .................... | .................... |
| V | 3266 | hCoV-19/Japan/PG-2526/2020\|EPI_ISL_690661\|2020-05 | .................... | .................... | .................... | .................... | .................... | .................... |
| V | 3267 | hCoV-19/Japan/PG-2527/2020\|EPI_ISL_690662\|2020-05 | .................... | .................... | .................... | .................... | .................... | .................... |
| V | 3268 | hCoV-19/Japan/PG-2528/2020\|EPI_ISL_690663\|2020-05 | .................... | .................... | .................... | .................... | .................... | .................... |
| V | 3269 | hCoV-19/Japan/PG-2529/2020\|EPI_ISL_690664\|2020-05 | .................... | .................... | .................... | .................... | .................... | .................... |
| V | 3270 | hCoV-19/Japan/PG-2530/2020\|EPI_ISL_690665\|2020-05 | .................... | .................... | .................... | .................... | .................... | .................... |
| V | 3271 | hCoV-19/Japan/PG-2531/2020\|EPI_ISL_690666\|2020-05 | .................... | .................... | .................... | .................... | .................... | .................... |
| V | 3272 | hCoV-19/Japan/PG-2532/2020\|EPI_ISL_690667\|2020-05 | .................... | .................... | .................... | .................... | .................... | .................... |
| V | 3273 | hCoV-19/Japan/PG-2533/2020\|EPI_ISL_690668\|2020-05 | .................... | .................... | .................... | .................... | .................... | .................... |
| V | 3274 | hCoV-19/Japan/PG-2534/2020\|EPI_ISL_690669\|2020-05 | .................... | .................... | .................... | .................... | .................... | .................... |
| V | 3275 | hCoV-19/Japan/PG-3647/2020\|EPI_ISL_691900\|2020-04 | .................... | .................... | .................... | .................... | .................... | .................... |
| V | 3276 | hCoV-19/Japan/PG-3648/2020\|EPI_ISL_691901\|2020-04 | .................... | .................... | .................... | .................... | .................... | .................... |
| V | 3277 | hCoV-19/Japan/PG-3649/2020\|EPI_ISL_691902\|2020-04 | .................... | .................... | .................... | .................... | .................... | .................... |
| V | 3278 | hCoV-19/Japan/PG-3650/2020\|EPI_ISL_691903\|2020-04 | .................... | .................... | .................... | .................... | .................... | .................... |
| V | 3279 | hCoV-19/Japan/PG-3651/2020\|EPI_ISL_691904\|2020-04 | .................... | .................... | .................... | .................... | .................... | .................... |
| V | 3280 | hCoV-19/Japan/PG-3652/2020\|EPI_ISL_691905\|2020-04 | .................... | .................... | .................... | .................... | .................... | .................... |
| V | 3281 | hCoV-19/Japan/PG-3653/2020\|EPI_ISL_691906\|2020-04 | .................... | .................... | .................... | .................... | .................... | .................... |
| V | 3282 | hCoV-19/Japan/PG-3654/2020\|EPI_ISL_691907\|2020-04 | .................... | .................... | .................... | .................... | .................... | .................... |
| V | 3283 | hCoV-19/Japan/PG-3655/2020\|EPI_ISL_691908\|2020-04 | .................... | .................... | .................... | .................... | .................... | .................... |
| V | 3284 | hCoV-19/Japan/PG-3656/2020\|EPI_ISL_691909\|2020-04 | .................... | .................... | .................... | .................... | .................... | .................... |
| V | 3285 | hCoV-19/Japan/PG-3657/2020\|EPI_ISL_691910\|2020-04 | .................... | .................... | .................... | .................... | .................... | .................... |
| V | 3286 | hCoV-19/Japan/PG-3658/2020\|EPI_ISL_691911\|2020-04 | .................... | .................... | .................... | .................... | .................... | .................... |
| V | 3287 | hCoV-19/Japan/PG-3659/2020\|EPI_ISL_691912\|2020-04 | .................... | .................... | .................... | .................... | .................... | .................... |
| V | 3288 | hCoV-19/Japan/PG-3660/2020\|EPI_ISL_691913\|2020-04 | .................... | .................... | .................... | .................... | .................... | .................... |
| V | 3289 | hCoV-19/Japan/PG-3661/2020\|EPI_ISL_691914\|2020-04 | .................... | .................... | .................... | .................... | .................... | .................... |
| V | 3290 | hCoV-19/Japan/PG-3662/2020\|EPI_ISL_691915\|2020-04 | .................... | .................... | .................... | .................... | .................... | .................... |
| V | 3291 | hCoV-19/Japan/PG-3663/2020\|EPI_ISL_691916\|2020-04 | .................... | .................... | .................... | .................... | .................... | .................... |
| V | 3292 | hCoV-19/Japan/PG-3664/2020\|EPI_ISL_691917\|2020-04 | .................... | .................... | .................... | .................... | .................... | .................... |
| V | 3293 | hCoV-19/Japan/PG-3665/2020\|EPI_ISL_691918\|2020-04 | .................... | .................... | .................... | .................... | .................... | .................... |
| V | 3294 | hCoV-19/Japan/PG-3666/2020\|EPI_ISL_691919\|2020-04 | .................... | .................... | .................... | .................... | .................... | .................... |
| V | 3295 | hCoV-19/Japan/PG-3708/2020\|EPI_ISL_691961\|2020-04 | .................... | .................... | .................... | .................... | .................... | .................... |
| V | 3296 | hCoV-19/Japan/PG-3709/2020\|EPI_ISL_691962\|2020-04 | .................... | .................... | .................... | .................... | .................... | .................... |
| V | 3297 | hCoV-19/Japan/PG-3710/2020\|EPI_ISL_691963\|2020-04 | .................... | .................... | .................... | .................... | .................... | .................... |
| V | 3298 | hCoV-19/Japan/PG-3711/2020\|EPI_ISL_691964\|2020-04 | .................... | .................... | .................... | .................... | .................... | .................... |
| V | 3299 | hCoV-19/Japan/PG-3717/2020\|EPI_ISL_691970\|2020-04 | .................... | .................... | .................... | .................... | .................... | .................... |
| V | 3300 | hCoV-19/Japan/PG-3718/2020\|EPI_ISL_691971\|2020-04 | .................... | .................... | .................... | .................... | .................... | .................... |
| V | 3301 | hCoV-19/Japan/PG-3719/2020\|EPI_ISL_691972\|2020-04 | .................... | .................... | .................... | .................... | .................... | .................... |
| V | 3302 | hCoV-19/Japan/PG-3720/2020\|EPI_ISL_691973\|2020-04 | .................... | .................... | .................... | .................... | .................... | .................... |
| V | 3303 | hCoV-19/Japan/PG-3721/2020\|EPI_ISL_691974\|2020-04 | .................... | .................... | .................... | .................... | .................... | .................... |
| V | 3304 | hCoV-19/Japan/PG-3722/2020\|EPI_ISL_691975\|2020-04 | .................... | .................... | .................... | .................... | .................... | .................... |
| V | 3305 | hCoV-19/Japan/PG-3723/2020\|EPI_ISL_691976\|2020-04 | .................... | .................... | .................... | .................... | .................... | .................... |
| V | 3306 | hCoV-19/Japan/PG-3724/2020\|EPI_ISL_691977\|2020-04 | .................... | .................... | .................... | .................... | .................... | .................... |
| V | 3307 | hCoV-19/Japan/PG-3725/2020\|EPI_ISL_691978\|2020-04 | .................... | .................... | .................... | .................... | .................... | .................... |
| V | 3308 | hCoV-19/Japan/PG-3726/2020\|EPI_ISL_691979\|2020-04 | .................... | .................... | .................... | .................... | .................... | .................... |
| V | 3309 | hCoV-19/Japan/PG-3727/2020\|EPI_ISL_691980\|2020-04 | .................... | .................... | .................... | .................... | .................... | .................... |
| V | 3310 | hCoV-19/Japan/PG-3728/2020\|EPI_ISL_691981\|2020-04 | .................... | .................... | .................... | .................... | .................... | .................... |
| V | 3311 | hCoV-19/Japan/PG-3729/2020\|EPI_ISL_691982\|2020-04 | .................... | .................... | .................... | .................... | .................... | .................... |
| V | 3312 | hCoV-19/Japan/PG-3730/2020\|EPI_ISL_691983\|2020-04 | .................... | .................... | .................... | .................... | .................... | .................... |
| V | 3313 | hCoV-19/Japan/PG-3731/2020\|EPI_ISL_691984\|2020-04 | .................... | .................... | .................... | .................... | .................... | .................... |
| V | 3314 | hCoV-19/Japan/PG-3732/2020\|EPI_ISL_691985\|2020-04 | .................... | .................... | .................... | .................... | .................... | .................... |
| V | 3315 | hCoV-19/Japan/PG-3733/2020\|EPI_ISL_691986\|2020-04 | .................... | .................... | .................... | .................... | .................... | .................... |
| V | 3316 | hCoV-19/Japan/PG-3734/2020\|EPI_ISL_691987\|2020-04 | .................... | .................... | .................... | .................... | .................... | .................... |
| V | 3317 | hCoV-19/Japan/PG-3735/2020\|EPI_ISL_691988\|2020-04 | .................... | .................... | .................... | .................... | .................... | .................... |
| V | 3318 | hCoV-19/Japan/PG-3736/2020\|EPI_ISL_691989\|2020-04 | .................... | .................... | .................... | .................... | .................... | .................... |
| V | 3319 | hCoV-19/Japan/PG-3737/2020\|EPI_ISL_691990\|2020-04 | .................... | .................... | .................... | .................... | .................... | .................... |
| V | 3320 | hCoV-19/Japan/PG-3738/2020\|EPI_ISL_691991\|2020-04 | .................... | .................... | .................... | .................... | .................... | .................... |
| V | 3321 | hCoV-19/Japan/PG-3739/2020\|EPI_ISL_691992\|2020-04 | .................... | .................... | .................... | .................... | .................... | .................... |
| V | 3322 | hCoV-19/Japan/PG-3740/2020\|EPI_ISL_691993\|2020-04 | .................... | .................... | .................... | .................... | .................... | .................... |
| V | 3323 | hCoV-19/Japan/PG-3741/2020\|EPI_ISL_691994\|2020-04 | .................... | .................... | .................... | .................... | .................... | .................... |
| V | 3324 | hCoV-19/Japan/PG-3742/2020\|EPI_ISL_691995\|2020-04 | .................... | .................... | .................... | .................... | .................... | .................... |
| V | 3325 | hCoV-19/Japan/PG-3743/2020\|EPI_ISL_691996\|2020-04 | .................... | .................... | .................... | .................... | .................... | .................... |
| V | 3326 | hCoV-19/Japan/PG-3744/2020\|EPI_ISL_691997\|2020-04 | .................... | .................... | .................... | .................... | .................... | .................... |
| V | 3327 | hCoV-19/Japan/PG-3745/2020\|EPI_ISL_691998\|2020-04 | .................... | .................... | .................... | .................... | .................... | .................... |
| V | 3328 | hCoV-19/Japan/PG-3746/2020\|EPI_ISL_691999\|2020-04 | .................... | .................... | .................... | .................... | .................... | .................... |
| V | 3329 | hCoV-19/Japan/PG-1090/2020\|EPI_ISL_684700\|2020-04 | .................... | .................... | .................... | .................... | .................... | .................... |
| V | 3330 | hCoV-19/Japan/PG-1091/2020\|EPI_ISL_684701\|2020-04 | .................... | .................... | .................... | .................... | .................... | .................... |
| V | 3331 | hCoV-19/Japan/PG-1092/2020\|EPI_ISL_684702\|2020-04 | .................... | .................... | .................... | .................... | .................... | .................... |
| V | 3332 | hCoV-19/Japan/PG-1093/2020\|EPI_ISL_684703\|2020-04 | .................... | .................... | .................... | .................... | .................... | .................... |
| V | 3333 | hCoV-19/Japan/PG-1094/2020\|EPI_ISL_684704\|2020-04 | .................... | .................... | .................... | .................... | .................... | .................... |
| V | 3334 | hCoV-19/Japan/PG-1095/2020\|EPI_ISL_684705\|2020-04 | .................... | .................... | .................... | .................... | .................... | .................... |
| V | 3335 | hCoV-19/Japan/PG-1096/2020\|EPI_ISL_684706\|2020-04 | .................... | .................... | .................... | .................... | .................... | .................... |
| V | 3336 | hCoV-19/Japan/PG-1097/2020\|EPI_ISL_684707\|2020-04 | .................... | .................... | .................... | .................... | .................... | .................... |
| V | 3337 | hCoV-19/Japan/PG-1098/2020\|EPI_ISL_684708\|2020-04 | .................... | .................... | .................... | .................... | .................... | .................... |
| V | 3338 | hCoV-19/Japan/PG-1099/2020\|EPI_ISL_684709\|2020-04 | .................... | .................... | .................... | .................... | .................... | .................... |

| | | | | | | | | |
|---|---|---|---|---|---|---|---|---|
| V | 3339 | hCoV-19/Japan/PG-7095/2020\|EPI_ISL_688200\|2020-08 | .................... | .................... | ..................... | .................... | .................... | .................... |
| V | 3340 | hCoV-19/Japan/PG-7096/2020\|EPI_ISL_688201\|2020-08 | .................... | .................... | ..................... | .................... | .................... | .................... |
| V | 3341 | hCoV-19/Japan/PG-7097/2020\|EPI_ISL_688202\|2020-08 | .................... | .................... | ..................... | .................... | .................... | .................... |
| V | 3342 | hCoV-19/Japan/PG-7098/2020\|EPI_ISL_688203\|2020-08 | .................... | .................... | ..................... | .................... | .................... | .................... |
| V | 3343 | hCoV-19/Japan/PG-7099/2020\|EPI_ISL_688204\|2020-08 | .................... | .................... | ..................... | .................... | .................... | .................... |
| V | 3344 | hCoV-19/Japan/PG-7100/2020\|EPI_ISL_688205\|2020-08 | .................... | .................... | ..................... | .................... | .................... | .................... |
| V | 3345 | hCoV-19/Japan/PG-7101/2020\|EPI_ISL_688206\|2020-08 | .................... | .................... | ..................... | .................... | .................... | .................... |
| V | 3346 | hCoV-19/Japan/PG-7102/2020\|EPI_ISL_688207\|2020-08 | .................... | .................... | ..................... | .................... | .................... | .................... |
| V | 3347 | hCoV-19/Japan/PG-7103/2020\|EPI_ISL_688208\|2020-08 | .................... | .................... | ..................... | .................... | .................... | .................... |
| V | 3348 | hCoV-19/Japan/PG-7104/2020\|EPI_ISL_688209\|2020-08 | .................... | .................... | ..................... | .................... | .................... | .................... |
| V | 3349 | hCoV-19/Japan/PG-7105/2020\|EPI_ISL_688210\|2020-08 | .................... | .................... | ..................... | .................... | .................... | .................... |
| V | 3350 | hCoV-19/Japan/PG-7106/2020\|EPI_ISL_688211\|2020-08 | .................... | .................... | ..................... | .................... | .................... | .................... |
| V | 3351 | hCoV-19/Japan/PG-7107/2020\|EPI_ISL_688212\|2020-08 | .................... | .................... | ..................... | .................... | .................... | .................... |
| V | 3352 | hCoV-19/Japan/PG-7108/2020\|EPI_ISL_688213\|2020-08 | .................... | .................... | ..................... | .................... | .................... | .................... |
| V | 3353 | hCoV-19/Japan/PG-7109/2020\|EPI_ISL_688214\|2020-08 | .................... | .................... | ..................... | .................... | .................... | .................... |
| V | 3354 | hCoV-19/Japan/PG-7110/2020\|EPI_ISL_688215\|2020-08 | .................... | .................... | ..................... | .................... | .................... | .................... |
| V | 3355 | hCoV-19/Japan/PG-7111/2020\|EPI_ISL_688216\|2020-08 | .................... | .................... | ..................... | .................... | .................... | .................... |
| V | 3356 | hCoV-19/Japan/PG-7112/2020\|EPI_ISL_688217\|2020-08 | .................... | .................... | ..................... | .................... | .................... | .................... |
| V | 3357 | hCoV-19/Japan/PG-7113/2020\|EPI_ISL_688218\|2020-08 | .................... | .................... | ..................... | .................... | .................... | .................... |
| V | 3358 | hCoV-19/Japan/PG-7114/2020\|EPI_ISL_688219\|2020-08 | .................... | .................... | ..................... | .................... | .................... | .................... |
| V | 3359 | hCoV-19/Japan/PG-7115/2020\|EPI_ISL_688220\|2020-08 | .................... | .................... | ..................... | .................... | .................... | .................... |
| V | 3360 | hCoV-19/Japan/PG-7116/2020\|EPI_ISL_688221\|2020-08 | .................... | .................... | ..................... | .................... | .................... | .................... |
| V | 3361 | hCoV-19/Japan/PG-7117/2020\|EPI_ISL_688222\|2020-08 | .................... | .................... | ..................... | .................... | .................... | .................... |
| V | 3362 | hCoV-19/Japan/PG-7118/2020\|EPI_ISL_688223\|2020-08 | .................... | .................... | ..................... | .................... | .................... | .................... |
| V | 3363 | hCoV-19/Japan/PG-7119/2020\|EPI_ISL_688224\|2020-08 | .................... | .................... | ..................... | .................... | .................... | .................... |
| V | 3364 | hCoV-19/Japan/PG-7120/2020\|EPI_ISL_688225\|2020-08 | .................... | .................... | ..................... | .................... | .................... | .................... |
| V | 3365 | hCoV-19/Japan/PG-7121/2020\|EPI_ISL_688226\|2020-08 | .................... | .................... | ..................... | .................... | .................... | .................... |
| V | 3366 | hCoV-19/Japan/PG-7122/2020\|EPI_ISL_688227\|2020-08 | .................... | .................... | ..................... | .................... | .................... | .................... |
| V | 3367 | hCoV-19/Japan/PG-7123/2020\|EPI_ISL_688228\|2020-08 | .................... | .................... | ..................... | .................... | .................... | .................... |
| V | 3368 | hCoV-19/Japan/PG-7124/2020\|EPI_ISL_688229\|2020-08 | .................... | .................... | ..................... | .................... | .................... | .................... |
| V | 3369 | hCoV-19/Japan/PG-7125/2020\|EPI_ISL_688230\|2020-08 | .................... | .................... | ..................... | .................... | .................... | .................... |
| V | 3370 | hCoV-19/Japan/PG-7126/2020\|EPI_ISL_688231\|2020-08 | .................... | .................... | ..................... | .................... | .................... | .................... |
| V | 3371 | hCoV-19/Japan/PG-7127/2020\|EPI_ISL_688232\|2020-08 | .................... | .................... | ..................... | .................... | .................... | .................... |
| V | 3372 | hCoV-19/Japan/PG-7128/2020\|EPI_ISL_688233\|2020-08 | .................... | .................... | ..................... | .................... | .................... | .................... |
| V | 3373 | hCoV-19/Japan/PG-7129/2020\|EPI_ISL_688234\|2020-08 | .................... | .................... | ..................... | .................... | .................... | .................... |
| V | 3374 | hCoV-19/Japan/PG-7130/2020\|EPI_ISL_688235\|2020-08 | .................... | .................... | ..................... | .................... | .................... | .................... |
| V | 3375 | hCoV-19/Japan/PG-7131/2020\|EPI_ISL_688236\|2020-08 | .................... | .................... | ..................... | .................... | .................... | .................... |
| V | 3376 | hCoV-19/Japan/PG-7175/2020\|EPI_ISL_688280\|2020-07 | .................... | .................... | ..................... | .................... | .................... | .................... |
| V | 3377 | hCoV-19/Japan/PG-7176/2020\|EPI_ISL_688281\|2020-08 | .................... | .................... | ..................... | .................... | .................... | .................... |
| V | 3378 | hCoV-19/Japan/PG-7177/2020\|EPI_ISL_688282\|2020-08 | .................... | .................... | ..................... | .................... | .................... | .................... |
| V | 3379 | hCoV-19/Japan/PG-7178/2020\|EPI_ISL_688283\|2020-08 | .................... | .................... | ..................... | .................... | .................... | .................... |
| V | 3380 | hCoV-19/Japan/PG-7179/2020\|EPI_ISL_688284\|2020-07 | .................... | .................... | ..................... | .................... | .................... | .................... |
| V | 3381 | hCoV-19/Japan/PG-7180/2020\|EPI_ISL_688285\|2020-07 | .................... | .................... | ..................... | .................... | .................... | .................... |
| V | 3382 | hCoV-19/Japan/PG-7181/2020\|EPI_ISL_688286\|2020-07 | .................... | .................... | ..................... | .................... | .................... | .................... |
| V | 3383 | hCoV-19/Japan/PG-7182/2020\|EPI_ISL_688287\|2020-07 | .................... | .................... | ..................... | .................... | .................... | .................... |
| V | 3384 | hCoV-19/Japan/PG-7184/2020\|EPI_ISL_688288\|2020-07 | .................... | .................... | ..................... | .................... | .................... | .................... |
| V | 3385 | hCoV-19/Japan/PG-7185/2020\|EPI_ISL_688289\|2020-07 | .................... | .................... | ..................... | .................... | .................... | .................... |
| V | 3386 | hCoV-19/Japan/PG-7186/2020\|EPI_ISL_688290\|2020-07 | .................... | .................... | ..................... | .................... | .................... | .................... |
| V | 3387 | hCoV-19/Japan/PG-7187/2020\|EPI_ISL_688291\|2020-07 | .................... | .................... | ..................... | .................... | .................... | .................... |
| V | 3388 | hCoV-19/Japan/PG-7188/2020\|EPI_ISL_688292\|2020-07 | .................... | .................... | ..................... | .................... | .................... | .................... |
| V | 3389 | hCoV-19/Japan/PG-7189/2020\|EPI_ISL_688293\|2020-07 | .................... | .................... | ..................... | .................... | .................... | .................... |
| V | 3390 | hCoV-19/Japan/PG-7190/2020\|EPI_ISL_688294\|2020-07 | .................... | .................... | ..................... | .................... | .................... | .................... |
| V | 3391 | hCoV-19/Japan/PG-7191/2020\|EPI_ISL_688295\|2020-07 | .................... | .................... | ..................... | .................... | .................... | .................... |
| V | 3392 | hCoV-19/Japan/PG-7192/2020\|EPI_ISL_688296\|2020-07 | .................... | .................... | ..................... | .................... | .................... | .................... |
| V | 3393 | hCoV-19/Japan/PG-7193/2020\|EPI_ISL_688297\|2020-07 | .................... | .................... | ..................... | .................... | .................... | .................... |
| V | 3394 | hCoV-19/Japan/PG-7194/2020\|EPI_ISL_688298\|2020-07 | .................... | .................... | ..................... | .................... | .................... | .................... |
| V | 3395 | hCoV-19/Japan/PG-7195/2020\|EPI_ISL_688299\|2020-07 | .................... | .................... | ..................... | .................... | .................... | .................... |
| V | 3396 | hCoV-19/Japan/PG-6394/2020\|EPI_ISL_689500\|2020-08 | .................... | .................... | ..................... | .................... | .................... | .................... |
| V | 3397 | hCoV-19/Japan/PG-6395/2020\|EPI_ISL_689501\|2020-08 | .................... | .................... | ..................... | .................... | .................... | .................... |
| V | 3398 | hCoV-19/Japan/PG-6396/2020\|EPI_ISL_689502\|2020-08 | .................... | .................... | ..................... | .................... | .................... | .................... |
| V | 3399 | hCoV-19/Japan/PG-6397/2020\|EPI_ISL_689503\|2020-08 | .................... | .................... | ..................... | .................... | .................... | .................... |
| V | 3400 | hCoV-19/Japan/PG-6398/2020\|EPI_ISL_689504\|2020-08 | .................... | .................... | ..................... | .................... | .................... | .................... |
| V | 3401 | hCoV-19/Japan/PG-6399/2020\|EPI_ISL_689505\|2020-08 | .................... | .................... | ..................... | .................... | .................... | .................... |
| V | 3402 | hCoV-19/Japan/PG-6400/2020\|EPI_ISL_689506\|2020-08 | .................... | .................... | ..................... | .................... | .................... | .................... |
| V | 3403 | hCoV-19/Japan/PG-6401/2020\|EPI_ISL_689507\|2020-08 | .................... | .................... | ..................... | .................... | .................... | .................... |
| V | 3404 | hCoV-19/Japan/PG-6402/2020\|EPI_ISL_689508\|2020-08 | .................... | .................... | ..................... | .................... | .................... | .................... |
| V | 3405 | hCoV-19/Japan/PG-6403/2020\|EPI_ISL_689509\|2020-08 | .................... | .................... | ..................... | .................... | .................... | .................... |
| V | 3406 | hCoV-19/Japan/PG-6404/2020\|EPI_ISL_689510\|2020-08 | .................... | .................... | ..................... | .................... | .................... | .................... |
| V | 3407 | hCoV-19/Japan/PG-6405/2020\|EPI_ISL_689511\|2020-08 | .................... | .................... | ..................... | .................... | .................... | .................... |
| V | 3408 | hCoV-19/Japan/PG-6406/2020\|EPI_ISL_689512\|2020-08 | .................... | .................... | ..................... | .................... | .................... | .................... |
| V | 3409 | hCoV-19/Japan/PG-6407/2020\|EPI_ISL_689513\|2020-08 | .................... | .................... | ..................... | .................... | .................... | .................... |
| V | 3410 | hCoV-19/Japan/PG-6408/2020\|EPI_ISL_689514\|2020-08 | .................... | .................... | ..................... | .................... | .................... | .................... |
| V | 3411 | hCoV-19/Japan/PG-6409/2020\|EPI_ISL_689515\|2020-08 | .................... | .................... | ..................... | .................... | .................... | .................... |
| V | 3412 | hCoV-19/Japan/PG-6410/2020\|EPI_ISL_689516\|2020-08 | .................... | .................... | ..................... | .................... | .................... | .................... |
| V | 3413 | hCoV-19/Japan/PG-6411/2020\|EPI_ISL_689517\|2020-07 | .................... | .................... | ..................... | .................... | .................... | .................... |
| V | 3414 | hCoV-19/Japan/PG-6412/2020\|EPI_ISL_689518\|2020-07 | .................... | .................... | ..................... | .................... | .................... | .................... |
| V | 3415 | hCoV-19/Japan/PG-6413/2020\|EPI_ISL_689519\|2020-07 | .................... | .................... | ..................... | .................... | .................... | .................... |
| V | 3416 | hCoV-19/Japan/PG-6414/2020\|EPI_ISL_689520\|2020-07 | .................... | .................... | ..................... | .................... | .................... | .................... |
| V | 3417 | hCoV-19/Japan/PG-6415/2020\|EPI_ISL_689521\|2020-07 | .................... | .................... | ..................... | .................... | .................... | .................... |
| V | 3418 | hCoV-19/Japan/PG-6416/2020\|EPI_ISL_689522\|2020-06 | .................... | .................... | ..................... | .................... | .................... | .................... |

| | | | | | | | |
|---|---|---|---|---|---|---|---|
| V | 3419 hCoV-19/Japan/PG-6417/2020\|EPI_ISL_689523\|2020-07 | .................... | .................... | ..................... | .................... | .................... | .................... |
| V | 3420 hCoV-19/Japan/PG-6418/2020\|EPI_ISL_689524\|2020-07 | .................... | .................... | ..................... | .................... | .................... | .................... |
| V | 3421 hCoV-19/Japan/PG-6419/2020\|EPI_ISL_689525\|2020-07 | .................... | .................... | ..................... | .................... | .................... | .................... |
| V | 3422 hCoV-19/Japan/PG-6420/2020\|EPI_ISL_689526\|2020-07 | .................... | .................... | ..................... | .................... | .................... | .................... |
| V | 3423 hCoV-19/Japan/PG-6421/2020\|EPI_ISL_689527\|2020-07 | .................... | .................... | ..................... | .................... | .................... | .................... |
| V | 3424 hCoV-19/Japan/PG-6422/2020\|EPI_ISL_689528\|2020-07 | .................... | .................... | ..................... | .................... | .................... | .................... |
| V | 3425 hCoV-19/Japan/PG-6423/2020\|EPI_ISL_689529\|2020-07 | .................... | .................... | ..................... | .................... | .................... | .................... |
| V | 3426 hCoV-19/Japan/PG-6424/2020\|EPI_ISL_689530\|2020-07 | .................... | .................... | ..................... | .................... | .................... | .................... |
| V | 3427 hCoV-19/Japan/PG-6425/2020\|EPI_ISL_689531\|2020-07 | .................... | .................... | ..................... | .................... | .................... | .................... |
| V | 3428 hCoV-19/Japan/PG-6426/2020\|EPI_ISL_689532\|2020-07 | .................... | .................... | ..................... | .................... | .................... | .................... |
| V | 3429 hCoV-19/Japan/PG-6427/2020\|EPI_ISL_689533\|2020-07 | .................... | .................... | ..................... | .................... | .................... | .................... |
| V | 3430 hCoV-19/Japan/PG-6428/2020\|EPI_ISL_689534\|2020-07 | .................... | .................... | ..................... | .................... | .................... | .................... |
| V | 3431 hCoV-19/Japan/PG-6429/2020\|EPI_ISL_689535\|2020-07 | .................... | .................... | ..................... | .................... | .................... | .................... |
| V | 3432 hCoV-19/Japan/PG-6430/2020\|EPI_ISL_689536\|2020-07 | .................... | .................... | ..................... | .................... | .................... | .................... |
| V | 3433 hCoV-19/Japan/PG-6431/2020\|EPI_ISL_689537\|2020-07 | .................... | .................... | ..................... | .................... | .................... | .................... |
| V | 3434 hCoV-19/Japan/PG-6432/2020\|EPI_ISL_689538\|2020-07 | .................... | .................... | ..................... | .................... | .................... | .................... |
| V | 3435 hCoV-19/Japan/PG-6433/2020\|EPI_ISL_689539\|2020-07 | .................... | .................... | ..................... | .................... | .................... | .................... |
| V | 3436 hCoV-19/Japan/PG-6434/2020\|EPI_ISL_689540\|2020-07 | .................... | .................... | ..................... | .................... | .................... | .................... |
| V | 3437 hCoV-19/Japan/PG-6435/2020\|EPI_ISL_689541\|2020-08 | .................... | .................... | ..................... | .................... | .................... | .................... |
| V | 3439 hCoV-19/Japan/PG-6437/2020\|EPI_ISL_689543\|2020-07 | .................... | .................... | ..................... | .................... | .................... | .................... |
| V | 3440 hCoV-19/Japan/PG-6438/2020\|EPI_ISL_689544\|2020-08 | .................... | .................... | ..................... | .................... | .................... | .................... |
| V | 3441 hCoV-19/Japan/PG-6439/2020\|EPI_ISL_689545\|2020-08 | .................... | .................... | ..................... | .................... | .................... | .................... |
| V | 3442 hCoV-19/Japan/PG-6440/2020\|EPI_ISL_689546\|2020-08 | .................... | .................... | ..................... | .................... | .................... | .................... |
| V | 3443 hCoV-19/Japan/PG-6441/2020\|EPI_ISL_689547\|2020-08 | .................... | .................... | ..................... | .................... | .................... | .................... |
| V | 3444 hCoV-19/Japan/PG-6442/2020\|EPI_ISL_689548\|2020-08 | .................... | .................... | ..................... | .................... | .................... | .................... |
| V | 3445 hCoV-19/Japan/PG-6443/2020\|EPI_ISL_689549\|2020-09 | .................... | .................... | ..................... | .................... | .................... | .................... |
| V | 3446 hCoV-19/Japan/PG-6444/2020\|EPI_ISL_689550\|2020-09 | .................... | .................... | ..................... | .................... | .................... | .................... |
| V | 3447 hCoV-19/Japan/PG-6446/2020\|EPI_ISL_689551\|2020-08 | .................... | .................... | ..................... | .................... | .................... | .................... |
| V | 3448 hCoV-19/Japan/PG-6447/2020\|EPI_ISL_689552\|2020-08 | .................... | .................... | ..................... | .................... | .................... | .................... |
| V | 3449 hCoV-19/Japan/PG-6448/2020\|EPI_ISL_689553\|2020-08 | .................... | .................... | ..................... | .................... | .................... | .................... |
| V | 3450 hCoV-19/Japan/PG-6449/2020\|EPI_ISL_689554\|2020-08 | .................... | .................... | ..................... | .................... | .................... | .................... |
| V | 3451 hCoV-19/Japan/PG-6450/2020\|EPI_ISL_689555\|2020-08 | .................... | .................... | ..................... | .................... | .................... | .................... |
| V | 3452 hCoV-19/Japan/PG-6451/2020\|EPI_ISL_689556\|2020-08 | .................... | .................... | ..................... | .................... | .................... | .................... |
| V | 3453 hCoV-19/Japan/PG-6452/2020\|EPI_ISL_689557\|2020-08 | .................... | .................... | ..................... | .................... | .................... | .................... |
| V | 3454 hCoV-19/Japan/PG-6453/2020\|EPI_ISL_689558\|2020-08 | .................... | .................... | ..................... | .................... | .................... | .................... |
| V | 3455 hCoV-19/Japan/PG-6454/2020\|EPI_ISL_689559\|2020-08 | .................... | .................... | ..................... | .................... | .................... | .................... |
| V | 3456 hCoV-19/Japan/PG-6455/2020\|EPI_ISL_689560\|2020-08 | .................... | .................... | ..................... | .................... | .................... | .................... |
| V | 3457 hCoV-19/Japan/PG-6456/2020\|EPI_ISL_689561\|2020-08 | .................... | .................... | ..................... | .................... | .................... | .................... |
| V | 3458 hCoV-19/Japan/PG-6457/2020\|EPI_ISL_689562\|2020-08 | .................... | .................... | ..................... | .................... | .................... | .................... |
| V | 3459 hCoV-19/Japan/PG-6458/2020\|EPI_ISL_689563\|2020-08 | .................... | .................... | ..................... | .................... | .................... | .................... |
| V | 3460 hCoV-19/Japan/PG-6459/2020\|EPI_ISL_689564\|2020-08 | .................... | .................... | ..................... | .................... | .................... | .................... |
| V | 3461 hCoV-19/Japan/PG-6460/2020\|EPI_ISL_689565\|2020-08 | .................... | .................... | ..................... | .................... | .................... | .................... |
| V | 3462 hCoV-19/Japan/PG-6461/2020\|EPI_ISL_689566\|2020-08 | .................... | .................... | ..................... | .................... | .................... | .................... |
| V | 3463 hCoV-19/Japan/PG-6462/2020\|EPI_ISL_689567\|2020-08 | .................... | .................... | ..................... | .................... | .................... | .................... |
| V | 3464 hCoV-19/Japan/PG-5397/2020\|EPI_ISL_690500\|2020-08 | .................... | .................... | ..................... | .................... | .................... | .................... |
| V | 3465 hCoV-19/Japan/PG-5398/2020\|EPI_ISL_690501\|2020-08 | .................... | .................... | ..................... | .................... | .................... | .................... |
| V | 3466 hCoV-19/Japan/PG-5399/2020\|EPI_ISL_690502\|2020-08 | .................... | .................... | ..................... | .................... | .................... | .................... |
| V | 3467 hCoV-19/Japan/PG-5400/2020\|EPI_ISL_690503\|2020-08 | .................... | .................... | ..................... | .................... | .................... | .................... |
| V | 3468 hCoV-19/Japan/PG-5401/2020\|EPI_ISL_690504\|2020-08 | .................... | .................... | ..................... | .................... | .................... | .................... |
| V | 3469 hCoV-19/Japan/PG-5402/2020\|EPI_ISL_690505\|2020-08 | .................... | .................... | ..................... | .................... | .................... | .................... |
| V | 3470 hCoV-19/Japan/PG-5403/2020\|EPI_ISL_690506\|2020-08 | .................... | .................... | ..................... | .................... | .................... | .................... |
| V | 3471 hCoV-19/Japan/PG-5404/2020\|EPI_ISL_690507\|2020-08 | .................... | .................... | ..................... | .................... | .................... | .................... |
| V | 3472 hCoV-19/Japan/PG-5405/2020\|EPI_ISL_690508\|2020-08 | .................... | .................... | ..................... | .................... | .................... | .................... |
| V | 3473 hCoV-19/Japan/PG-5406/2020\|EPI_ISL_690509\|2020-08 | .................... | .................... | ..................... | .................... | .................... | .................... |
| V | 3474 hCoV-19/Japan/PG-5411/2020\|EPI_ISL_690514\|2020-08 | .................... | .................... | ..................... | .................... | .................... | .................... |
| V | 3475 hCoV-19/Japan/PG-5447/2020\|EPI_ISL_690550\|2020-08 | .................... | .................... | ..................... | .................... | .................... | .................... |
| V | 3476 hCoV-19/Japan/PG-5448/2020\|EPI_ISL_690551\|2020-08 | .................... | .................... | ..................... | .................... | .................... | .................... |
| V | 3477 hCoV-19/Japan/PG-5449/2020\|EPI_ISL_690552\|2020-08 | .................... | .................... | ..................... | .................... | .................... | .................... |
| V | 3478 hCoV-19/Japan/PG-5450/2020\|EPI_ISL_690553\|2020-08 | .................... | .................... | ..................... | .................... | .................... | .................... |
| V | 3479 hCoV-19/Japan/PG-5451/2020\|EPI_ISL_690554\|2020-08 | .................... | .................... | ..................... | .................... | .................... | .................... |
| V | 3480 hCoV-19/Japan/PG-5452/2020\|EPI_ISL_690555\|2020-08 | .................... | .................... | ..................... | .................... | .................... | .................... |
| V | 3481 hCoV-19/Japan/PG-5453/2020\|EPI_ISL_690556\|2020-08 | .................... | .................... | ..................... | .................... | .................... | .................... |
| V | 3482 hCoV-19/Japan/PG-5456/2020\|EPI_ISL_690559\|2020-08 | .................... | .................... | ..................... | .................... | .................... | .................... |
| V | 3483 hCoV-19/Japan/PG-5457/2020\|EPI_ISL_690560\|2020-08 | .................... | .................... | ..................... | .................... | .................... | .................... |
| V | 3484 hCoV-19/Japan/PG-5458/2020\|EPI_ISL_690561\|2020-08 | .................... | .................... | ..................... | .................... | .................... | .................... |
| V | 3485 hCoV-19/Japan/PG-5459/2020\|EPI_ISL_690562\|2020-08 | .................... | .................... | ..................... | .................... | .................... | .................... |
| V | 3486 hCoV-19/Japan/PG-5460/2020\|EPI_ISL_690563\|2020-08 | .................... | .................... | ..................... | .................... | .................... | .................... |
| V | 3488 hCoV-19/Japan/PG-5462/2020\|EPI_ISL_690565\|2020-08 | .................... | .................... | ..................... | .................... | .................... | .................... |
| V | 3489 hCoV-19/Japan/PG-5463/2020\|EPI_ISL_690566\|2020-08 | .................... | .................... | ..................... | .................... | .................... | .................... |
| V | 3490 hCoV-19/Japan/PG-5464/2020\|EPI_ISL_690567\|2020-08 | .................... | .................... | ..................... | .................... | .................... | .................... |
| V | 3491 hCoV-19/Japan/PG-5465/2020\|EPI_ISL_690568\|2020-08 | .................... | .................... | ..................... | .................... | .................... | .................... |
| V | 3492 hCoV-19/Japan/PG-5466/2020\|EPI_ISL_690569\|2020-08 | .................... | .................... | ..................... | .................... | .................... | .................... |
| V | 3493 hCoV-19/Japan/PG-5467/2020\|EPI_ISL_690570\|2020-08 | .................... | .................... | ..................... | .................... | .................... | .................... |
| V | 3494 hCoV-19/Japan/PG-5468/2020\|EPI_ISL_690571\|2020-08 | .................... | .................... | ..................... | .................... | .................... | .................... |
| V | 3495 hCoV-19/Japan/PG-5469/2020\|EPI_ISL_690572\|2020-08 | .................... | .................... | ..................... | .................... | .................... | .................... |
| V | 3496 hCoV-19/Japan/PG-5470/2020\|EPI_ISL_690573\|2020-07 | .................... | .................... | ..................... | .................... | .................... | .................... |
| V | 3497 hCoV-19/Japan/PG-5471/2020\|EPI_ISL_690574\|2020-07 | .................... | .................... | ..................... | .................... | .................... | .................... |
| V | 3498 hCoV-19/Japan/PG-5472/2020\|EPI_ISL_690575\|2020-08 | .................... | .................... | ..................... | .................... | .................... | .................... |
| V | 3499 hCoV-19/Japan/PG-5473/2020\|EPI_ISL_690576\|2020-08 | .................... | .................... | ..................... | .................... | .................... | .................... |
| V | 3500 hCoV-19/Japan/PG-5474/2020\|EPI_ISL_690577\|2020-08 | .................... | .................... | ..................... | .................... | .................... | .................... |

| | | | | | | | |
|---|---|---|---|---|---|---|---|
| V | 3501 hCoV-19/Japan/PG-5475/2020\|EPI_ISL_690578\|2020-08 | ···················· | ···················· | ···················· | ···················· | ···················· | ···················· |
| V | 3502 hCoV-19/Japan/PG-5476/2020\|EPI_ISL_690579\|2020-08 | ···················· | ···················· | ···················· | ···················· | ···················· | ···················· |
| V | 3503 hCoV-19/Japan/PG-5477/2020\|EPI_ISL_690580\|2020-08 | ···················· | ···················· | ···················· | ···················· | ···················· | ···················· |
| V | 3504 hCoV-19/Japan/PG-5478/2020\|EPI_ISL_690581\|2020-08 | ···················· | ···················· | ···················· | ···················· | ···················· | ···················· |
| V | 3505 hCoV-19/Japan/PG-5479/2020\|EPI_ISL_690582\|2020-08 | ···················· | ···················· | ···················· | ···················· | ···················· | ···················· |
| V | 3506 hCoV-19/Japan/PG-5480/2020\|EPI_ISL_690583\|2020-08 | ···················· | ···················· | ···················· | ···················· | ···················· | ···················· |
| V | 3507 hCoV-19/Japan/PG-5481/2020\|EPI_ISL_690584\|2020-08 | ···················· | ···················· | ···················· | ···················· | ···················· | ···················· |
| V | 3508 hCoV-19/Japan/PG-5482/2020\|EPI_ISL_690585\|2020-08 | ···················· | ···················· | ···················· | ···················· | ···················· | ···················· |
| V | 3509 hCoV-19/Japan/PG-5484/2020\|EPI_ISL_690586\|2020-08 | ···················· | ···················· | ···················· | ···················· | ···················· | ···················· |
| V | 3510 hCoV-19/Japan/PG-5485/2020\|EPI_ISL_690587\|2020-08 | ···················· | ···················· | ···················· | ···················· | ···················· | ···················· |
| V | 3511 hCoV-19/Japan/PG-5486/2020\|EPI_ISL_690588\|2020-08 | ···················· | ···················· | ···················· | ···················· | ···················· | ···················· |
| V | 3512 hCoV-19/Japan/PG-5487/2020\|EPI_ISL_690589\|2020-08 | ···················· | ···················· | ···················· | ···················· | ···················· | ···················· |
| V | 3513 hCoV-19/Japan/PG-5488/2020\|EPI_ISL_690590\|2020-08 | ···················· | ···················· | ···················· | ···················· | ···················· | ···················· |
| V | 3514 hCoV-19/Japan/PG-5489/2020\|EPI_ISL_690591\|2020-08 | ···················· | ···················· | ···················· | ···················· | ···················· | ···················· |
| V | 3515 hCoV-19/Japan/PG-5490/2020\|EPI_ISL_690592\|2020-08 | ···················· | ···················· | ···················· | ···················· | ···················· | ···················· |
| V | 3516 hCoV-19/Japan/PG-5491/2020\|EPI_ISL_690593\|2020-08 | ···················· | ···················· | ···················· | ···················· | ···················· | ···················· |
| V | 3517 hCoV-19/Japan/PG-5492/2020\|EPI_ISL_690594\|2020-08 | ···················· | ···················· | ···················· | ···················· | ···················· | ···················· |
| V | 3518 hCoV-19/Japan/PG-5493/2020\|EPI_ISL_690595\|2020-08 | ···················· | ···················· | ···················· | ···················· | ···················· | ···················· |
| V | 3519 hCoV-19/Japan/PG-5494/2020\|EPI_ISL_690596\|2020-08 | ···················· | ···················· | ···················· | ···················· | ···················· | ···················· |
| V | 3520 hCoV-19/Japan/PG-5495/2020\|EPI_ISL_690597\|2020-08 | ···················· | ···················· | ···················· | ···················· | ···················· | ···················· |
| V | 3521 hCoV-19/Japan/PG-5496/2020\|EPI_ISL_690598\|2020-08 | ···················· | ···················· | ···················· | ···················· | ···················· | ···················· |
| V | 3522 hCoV-19/Japan/PG-5497/2020\|EPI_ISL_690599\|2020-08 | ···················· | ···················· | ···················· | ···················· | ···················· | ···················· |
| V | 3523 hCoV-19/Japan/PG-3544/2020\|EPI_ISL_691800\|2020-06 | ···················· | ···················· | ···················· | ···················· | ···················· | ···················· |
| V | 3524 hCoV-19/Japan/PG-3545/2020\|EPI_ISL_691801\|2020-06 | ···················· | ···················· | ···················· | ···················· | ···················· | ···················· |
| V | 3525 hCoV-19/Japan/PG-3546/2020\|EPI_ISL_691802\|2020-06 | ···················· | ···················· | ···················· | ···················· | ···················· | ···················· |
| V | 3526 hCoV-19/Japan/PG-3547/2020\|EPI_ISL_691803\|2020-06 | ···················· | ···················· | ···················· | ···················· | ···················· | ···················· |
| V | 3527 hCoV-19/Japan/PG-3548/2020\|EPI_ISL_691804\|2020-06 | ···················· | ···················· | ···················· | ···················· | ···················· | ···················· |
| V | 3528 hCoV-19/Japan/PG-3549/2020\|EPI_ISL_691805\|2020-06 | ···················· | ···················· | ···················· | ···················· | ···················· | ···················· |
| V | 3529 hCoV-19/Japan/PG-3550/2020\|EPI_ISL_691806\|2020-06 | ···················· | ···················· | ···················· | ···················· | ···················· | ···················· |
| V | 3530 hCoV-19/Japan/PG-3551/2020\|EPI_ISL_691807\|2020-06 | ···················· | ···················· | ···················· | ···················· | ···················· | ···················· |
| V | 3531 hCoV-19/Japan/PG-3552/2020\|EPI_ISL_691808\|2020-06 | ···················· | ···················· | ···················· | ···················· | ···················· | ···················· |
| V | 3532 hCoV-19/Japan/PG-3553/2020\|EPI_ISL_691809\|2020-06 | ···················· | ···················· | ···················· | ···················· | ···················· | ···················· |
| V | 3533 hCoV-19/Japan/PG-3554/2020\|EPI_ISL_691810\|2020-06 | ···················· | ···················· | ···················· | ···················· | ···················· | ···················· |
| V | 3534 hCoV-19/Japan/PG-3555/2020\|EPI_ISL_691811\|2020-06 | ···················· | ···················· | ···················· | ···················· | ···················· | ···················· |
| V | 3535 hCoV-19/Japan/PG-3556/2020\|EPI_ISL_691812\|2020-06 | ···················· | ···················· | ···················· | ···················· | ···················· | ···················· |
| V | 3536 hCoV-19/Japan/PG-3557/2020\|EPI_ISL_691813\|2020-06 | ···················· | ···················· | ···················· | ···················· | ···················· | ···················· |
| V | 3537 hCoV-19/Japan/PG-3558/2020\|EPI_ISL_691814\|2020-06 | ···················· | ···················· | ···················· | ···················· | ···················· | ···················· |
| V | 3538 hCoV-19/Japan/PG-3559/2020\|EPI_ISL_691815\|2020-06 | ···················· | ···················· | ···················· | ···················· | ···················· | ···················· |
| V | 3539 hCoV-19/Japan/PG-3560/2020\|EPI_ISL_691816\|2020-06 | ···················· | ···················· | ···················· | ···················· | ···················· | ···················· |
| V | 3540 hCoV-19/Japan/PG-3561/2020\|EPI_ISL_691817\|2020-06 | ···················· | ···················· | ···················· | ···················· | ···················· | ···················· |
| V | 3541 hCoV-19/Japan/PG-3562/2020\|EPI_ISL_691818\|2020-06 | ···················· | ···················· | ···················· | ···················· | ···················· | ···················· |
| V | 3542 hCoV-19/Japan/PG-3563/2020\|EPI_ISL_691819\|2020-06 | ···················· | ···················· | ···················· | ···················· | ···················· | ···················· |
| V | 3543 hCoV-19/Japan/PG-3564/2020\|EPI_ISL_691820\|2020-06 | ···················· | ···················· | ···················· | ···················· | ···················· | ···················· |
| V | 3544 hCoV-19/Japan/PG-3565/2020\|EPI_ISL_691821\|2020-06 | ···················· | ···················· | ···················· | ···················· | ···················· | ···················· |
| V | 3545 hCoV-19/Japan/PG-3566/2020\|EPI_ISL_691822\|2020-06 | ···················· | ···················· | ···················· | ···················· | ···················· | ···················· |
| V | 3546 hCoV-19/Japan/PG-3567/2020\|EPI_ISL_691823\|2020-06 | ···················· | ···················· | ···················· | ···················· | ···················· | ···················· |
| V | 3547 hCoV-19/Japan/PG-3568/2020\|EPI_ISL_691824\|2020-06 | ···················· | ···················· | ···················· | ···················· | ···················· | ···················· |
| V | 3548 hCoV-19/Japan/PG-3569/2020\|EPI_ISL_691825\|2020-06 | ···················· | ···················· | ···················· | ···················· | ···················· | ···················· |
| V | 3549 hCoV-19/Japan/PG-3570/2020\|EPI_ISL_691826\|2020-06 | ···················· | ···················· | ···················· | ···················· | ···················· | ···················· |
| V | 3550 hCoV-19/Japan/PG-3571/2020\|EPI_ISL_691827\|2020-06 | ···················· | ···················· | ···················· | ···················· | ···················· | ···················· |
| V | 3551 hCoV-19/Japan/PG-3572/2020\|EPI_ISL_691828\|2020-06 | ···················· | ···················· | ···················· | ···················· | ···················· | ···················· |
| V | 3552 hCoV-19/Japan/PG-3573/2020\|EPI_ISL_691829\|2020-06 | ···················· | ···················· | ···················· | ···················· | ···················· | ···················· |
| V | 3553 hCoV-19/Japan/PG-3574/2020\|EPI_ISL_691830\|2020-06 | ···················· | ···················· | ···················· | ···················· | ···················· | ···················· |
| V | 3554 hCoV-19/Japan/PG-3575/2020\|EPI_ISL_691831\|2020-06 | ···················· | ···················· | ···················· | ···················· | ···················· | ···················· |
| V | 3555 hCoV-19/Japan/PG-3576/2020\|EPI_ISL_691832\|2020-06 | ···················· | ···················· | ···················· | ···················· | ···················· | ···················· |
| V | 3556 hCoV-19/Japan/PG-3577/2020\|EPI_ISL_691833\|2020-06 | ···················· | ···················· | ···················· | ···················· | ···················· | ···················· |
| V | 3557 hCoV-19/Japan/PG-3578/2020\|EPI_ISL_691834\|2020-06 | ···················· | ···················· | ···················· | ···················· | ···················· | ···················· |
| V | 3558 hCoV-19/Japan/PG-3579/2020\|EPI_ISL_691835\|2020-06 | ···················· | ···················· | ···················· | ···················· | ···················· | ···················· |
| V | 3559 hCoV-19/Japan/PG-3580/2020\|EPI_ISL_691836\|2020-06 | ···················· | ···················· | ···················· | ···················· | ···················· | ···················· |
| V | 3560 hCoV-19/Japan/PG-3581/2020\|EPI_ISL_691837\|2020-06 | ···················· | ···················· | ···················· | ···················· | ···················· | ···················· |
| V | 3561 hCoV-19/Japan/PG-3582/2020\|EPI_ISL_691838\|2020-06 | ···················· | ···················· | ···················· | ···················· | ·········T·········· | ···················· |
| V | 3562 hCoV-19/Japan/PG-3583/2020\|EPI_ISL_691839\|2020-06 | ···················· | ···················· | ···················· | ···················· | ···················· | ···················· |
| V | 3563 hCoV-19/Japan/PG-3592/2020\|EPI_ISL_691846\|2020-07 | ···················· | ···················· | ···················· | ···················· | ···················· | ···················· |
| V | 3564 hCoV-19/Japan/PG-3593/2020\|EPI_ISL_691847\|2020-07 | ···················· | ···················· | ···················· | ···················· | ···················· | ···················· |
| V | 3565 hCoV-19/Japan/PG-3627/2020\|EPI_ISL_691880\|2020-04 | ···················· | ···················· | ···················· | ···················· | ···················· | ···················· |
| V | 3566 hCoV-19/Japan/PG-3628/2020\|EPI_ISL_691881\|2020-04 | ···················· | ···················· | ···················· | ···················· | ···················· | ···················· |
| V | 3567 hCoV-19/Japan/PG-3629/2020\|EPI_ISL_691882\|2020-04 | ···················· | ···················· | ···················· | ···················· | ···················· | ···················· |
| V | 3568 hCoV-19/Japan/PG-3630/2020\|EPI_ISL_691883\|2020-04 | ···················· | ···················· | ···················· | ···················· | ···················· | ···················· |
| V | 3569 hCoV-19/Japan/PG-3631/2020\|EPI_ISL_691884\|2020-04 | ···················· | ···················· | ···················· | ···················· | ···················· | ···················· |
| V | 3570 hCoV-19/Japan/PG-3632/2020\|EPI_ISL_691885\|2020-04 | ···················· | ···················· | ···················· | ···················· | ···················· | ···················· |
| V | 3571 hCoV-19/Japan/PG-3633/2020\|EPI_ISL_691886\|2020-04 | ···················· | ···················· | ···················· | ···················· | ···················· | ···················· |
| V | 3572 hCoV-19/Japan/PG-3634/2020\|EPI_ISL_691887\|2020-04 | ···················· | ···················· | ···················· | ···················· | ···················· | ···················· |
| V | 3573 hCoV-19/Japan/PG-3635/2020\|EPI_ISL_691888\|2020-04 | ···················· | ···················· | ···················· | ···················· | ···················· | ···················· |
| V | 3574 hCoV-19/Japan/PG-3636/2020\|EPI_ISL_691889\|2020-04 | ···················· | ···················· | ···················· | ···················· | ···················· | ···················· |
| V | 3575 hCoV-19/Japan/PG-3637/2020\|EPI_ISL_691890\|2020-04 | ···················· | ···················· | ···················· | ···················· | ···················· | ···················· |
| V | 3576 hCoV-19/Japan/PG-3638/2020\|EPI_ISL_691891\|2020-04 | ···················· | ···················· | ···················· | ···················· | ···················· | ···················· |
| V | 3577 hCoV-19/Japan/PG-3639/2020\|EPI_ISL_691892\|2020-04 | ···················· | ···················· | ···················· | ···················· | ···················· | ···················· |
| V | 3578 hCoV-19/Japan/PG-3640/2020\|EPI_ISL_691893\|2020-04 | ···················· | ···················· | ···················· | ···················· | ···················· | ···················· |
| V | 3579 hCoV-19/Japan/PG-3641/2020\|EPI_ISL_691894\|2020-04 | ···················· | ···················· | ···················· | ···················· | ···················· | ···················· |
| V | 3580 hCoV-19/Japan/PG-3642/2020\|EPI_ISL_691895\|2020-04 | ···················· | ···················· | ···················· | ···················· | ···················· | ···················· |

| | | | | | | | | |
|---|---|---|---|---|---|---|---|---|
| V | 3581 | hCoV-19/Japan/PG-3643/2020\|EPI_ISL_691896\|2020-04 | .................... | .................... | ..................... | .................... | .................... | .................... |
| V | 3582 | hCoV-19/Japan/PG-3644/2020\|EPI_ISL_691897\|2020-04 | .................... | .................... | ..................... | .................... | .................... | .................... |
| V | 3583 | hCoV-19/Japan/PG-3645/2020\|EPI_ISL_691898\|2020-04 | .................... | .................... | ..................... | .................... | .................... | .................... |
| V | 3584 | hCoV-19/Japan/PG-3646/2020\|EPI_ISL_691899\|2020-04 | .................... | .................... | ..................... | .................... | .................... | .................... |
| V | 3585 | hCoV-19/Japan/PG-1018/2020\|EPI_ISL_684628\|2020-03 | .................... | .................... | ..................... | .................... | .................... | .................... |
| V | 3586 | hCoV-19/Japan/PG-1019/2020\|EPI_ISL_684629\|2020-03 | .................... | .................... | ..................... | .................... | .................... | .................... |
| V | 3587 | hCoV-19/Japan/PG-1020/2020\|EPI_ISL_684630\|2020-03 | .................... | .................... | ..................... | .................... | .................... | .................... |
| V | 3588 | hCoV-19/Japan/PG-1021/2020\|EPI_ISL_684631\|2020-03 | .................... | .................... | ..................... | .................... | .................... | .................... |
| V | 3589 | hCoV-19/Japan/PG-1022/2020\|EPI_ISL_684632\|2020-04 | .................... | .................... | ..................... | .................... | .................... | .................... |
| V | 3590 | hCoV-19/Japan/PG-1023/2020\|EPI_ISL_684633\|2020-04 | .................... | .................... | ..................... | .................... | .................... | .................... |
| V | 3591 | hCoV-19/Japan/PG-1024/2020\|EPI_ISL_684634\|2020-04 | .................... | .................... | ..................... | .................... | .................... | .................... |
| V | 3592 | hCoV-19/Japan/PG-1025/2020\|EPI_ISL_684635\|2020-04 | .................... | .................... | ..................... | .................... | .................... | .................... |
| V | 3593 | hCoV-19/Japan/PG-1026/2020\|EPI_ISL_684636\|2020-04 | .................... | .................... | ..................... | .................... | .................... | .................... |
| V | 3594 | hCoV-19/Japan/PG-1027/2020\|EPI_ISL_684637\|2020-04 | .................... | .................... | ..................... | .................... | .................... | .................... |
| V | 3595 | hCoV-19/Japan/PG-1028/2020\|EPI_ISL_684638\|2020-04 | .................... | .................... | ..................... | .................... | .................... | .................... |
| V | 3596 | hCoV-19/Japan/PG-1030/2020\|EPI_ISL_684640\|2020-04 | .................... | .................... | ..................... | .................... | .................... | .................... |
| V | 3597 | hCoV-19/Japan/PG-1031/2020\|EPI_ISL_684641\|2020-04 | .................... | .................... | ..................... | .................... | .................... | .................... |
| V | 3598 | hCoV-19/Japan/PG-1032/2020\|EPI_ISL_684642\|2020-04 | .................... | .................... | ..................... | .................... | .................... | .................... |
| V | 3599 | hCoV-19/Japan/PG-1033/2020\|EPI_ISL_684643\|2020-04 | .................... | .................... | ..................... | .................... | .................... | .................... |
| V | 3600 | hCoV-19/Japan/PG-1034/2020\|EPI_ISL_684644\|2020-04 | .................... | .................... | ..................... | .................... | .................... | .................... |
| V | 3601 | hCoV-19/Japan/PG-1035/2020\|EPI_ISL_684645\|2020-04 | .................... | .................... | ..................... | .................... | .................... | .................... |
| V | 3602 | hCoV-19/Japan/PG-1036/2020\|EPI_ISL_684646\|2020-04 | .................... | .................... | ..................... | .................... | .................... | .................... |
| V | 3603 | hCoV-19/Japan/PG-1037/2020\|EPI_ISL_684647\|2020-04 | .................... | .................... | ..................... | .................... | .................... | .................... |
| V | 3604 | hCoV-19/Japan/PG-1038/2020\|EPI_ISL_684648\|2020-04 | .................... | .................... | ..................... | .................... | .................... | .................... |
| V | 3605 | hCoV-19/Japan/PG-1039/2020\|EPI_ISL_684649\|2020-04 | .................... | .................... | ..................... | .................... | .................... | .................... |
| V | 3606 | hCoV-19/Japan/PG-1040/2020\|EPI_ISL_684650\|2020-04 | .................... | .................... | ..................... | .................... | .................... | .................... |
| V | 3607 | hCoV-19/Japan/PG-1041/2020\|EPI_ISL_684651\|2020-04 | .................... | .................... | ..................... | .................... | .................... | .................... |
| V | 3608 | hCoV-19/Japan/PG-1042/2020\|EPI_ISL_684652\|2020-04 | .................... | .................... | ..................... | .................... | .................... | .................... |
| V | 3609 | hCoV-19/Japan/PG-1043/2020\|EPI_ISL_684653\|2020-04 | .................... | .................... | ..................... | .................... | .................... | .................... |
| V | 3610 | hCoV-19/Japan/PG-1044/2020\|EPI_ISL_684654\|2020-04 | .................... | .................... | ..................... | .................... | .................... | .................... |
| V | 3611 | hCoV-19/Japan/PG-1045/2020\|EPI_ISL_684655\|2020-04 | .................... | .................... | ..................... | .................... | .................... | .................... |
| V | 3612 | hCoV-19/Japan/PG-1046/2020\|EPI_ISL_684656\|2020-04 | .................... | .................... | ..................... | .................... | .................... | .................... |
| V | 3613 | hCoV-19/Japan/PG-1047/2020\|EPI_ISL_684657\|2020-04 | .................... | .................... | ..................... | .................... | .................... | .................... |
| V | 3614 | hCoV-19/Japan/PG-1048/2020\|EPI_ISL_684658\|2020-04 | .................... | .................... | ..................... | .................... | .................... | .................... |
| V | 3615 | hCoV-19/Japan/PG-1049/2020\|EPI_ISL_684659\|2020-04 | .................... | .................... | ..................... | .................... | .................... | .................... |
| V | 3616 | hCoV-19/Japan/PG-1050/2020\|EPI_ISL_684660\|2020-04 | .................... | .................... | ..................... | .................... | .................... | .................... |
| V | 3617 | hCoV-19/Japan/PG-1051/2020\|EPI_ISL_684661\|2020-04 | .................... | .................... | ..................... | .................... | .................... | .................... |
| V | 3618 | hCoV-19/Japan/PG-1052/2020\|EPI_ISL_684662\|2020-04 | .................... | .................... | ..................... | .................... | .................... | .................... |
| V | 3619 | hCoV-19/Japan/PG-1053/2020\|EPI_ISL_684663\|2020-04 | .................... | .................... | ..................... | .................... | .................... | .................... |
| V | 3620 | hCoV-19/Japan/PG-1054/2020\|EPI_ISL_684664\|2020-04 | .................... | .................... | ..................... | .................... | .................... | .................... |
| V | 3621 | hCoV-19/Japan/PG-1055/2020\|EPI_ISL_684665\|2020-04 | .................... | .................... | ..................... | .................... | .................... | .................... |
| V | 3622 | hCoV-19/Japan/PG-1056/2020\|EPI_ISL_684666\|2020-04 | .................... | .................... | ..................... | .................... | .................... | .................... |
| V | 3623 | hCoV-19/Japan/PG-1057/2020\|EPI_ISL_684667\|2020-04 | .................... | .................... | ..................... | .................... | .................... | .................... |
| V | 3624 | hCoV-19/Japan/PG-1058/2020\|EPI_ISL_684668\|2020-04 | .................... | .................... | ..................... | .................... | .................... | .................... |
| V | 3625 | hCoV-19/Japan/PG-1059/2020\|EPI_ISL_684669\|2020-04 | .................... | .................... | ..................... | .................... | .................... | .................... |
| V | 3626 | hCoV-19/Japan/PG-1060/2020\|EPI_ISL_684670\|2020-04 | .................... | .................... | ..................... | .................... | .................... | .................... |
| V | 3627 | hCoV-19/Japan/PG-1061/2020\|EPI_ISL_684671\|2020-04 | .................... | .................... | ..................... | .................... | .................... | .................... |
| V | 3628 | hCoV-19/Japan/PG-1062/2020\|EPI_ISL_684672\|2020-04 | .................... | .................... | ..................... | .................... | .................... | .................... |
| V | 3629 | hCoV-19/Japan/PG-1063/2020\|EPI_ISL_684673\|2020-04 | .................... | .................... | ..................... | .................... | .................... | .................... |
| V | 3630 | hCoV-19/Japan/PG-1064/2020\|EPI_ISL_684674\|2020-04 | .................... | .................... | ..................... | .................... | .................... | .................... |
| V | 3632 | hCoV-19/Japan/PG-2355/2020\|EPI_ISL_685960\|2020-04 | .................... | .................... | ..................... | .................... | .................... | .................... |
| V | 3633 | hCoV-19/Japan/PG-2356/2020\|EPI_ISL_685961\|2020-04 | .................... | .................... | ..................... | .................... | .................... | .................... |
| V | 3634 | hCoV-19/Japan/PG-2357/2020\|EPI_ISL_685962\|2020-03 | .................... | .................... | ..................... | .................... | .................... | .................... |
| V | 3635 | hCoV-19/Japan/PG-2358/2020\|EPI_ISL_685963\|2020-05 | .................... | .................... | ..................... | .................... | .................... | .................... |
| V | 3637 | hCoV-19/Japan/PG-2360/2020\|EPI_ISL_685965\|2020-05 | .................... | .................... | ..................... | .................... | .................... | .................... |
| V | 3638 | hCoV-19/Japan/PG-2361/2020\|EPI_ISL_685966\|2020-05 | .................... | .................... | ..................... | .................... | .................... | .................... |
| V | 3639 | hCoV-19/Japan/PG-2362/2020\|EPI_ISL_685967\|2020-05 | .................... | .................... | ..................... | .................... | .................... | .................... |
| V | 3640 | hCoV-19/Japan/PG-2363/2020\|EPI_ISL_685968\|2020-05 | .................... | .................... | ..................... | .................... | .................... | .................... |
| V | 3641 | hCoV-19/Japan/PG-2364/2020\|EPI_ISL_685969\|2020-05 | .................... | .................... | ..................... | .................... | .................... | .................... |
| V | 3642 | hCoV-19/Japan/PG-2365/2020\|EPI_ISL_685970\|2020-05 | .................... | .................... | ..................... | .................... | .................... | .................... |
| V | 3643 | hCoV-19/Japan/PG-2366/2020\|EPI_ISL_685971\|2020-05 | .................... | .................... | ..................... | .................... | .................... | .................... |
| V | 3644 | hCoV-19/Japan/PG-2367/2020\|EPI_ISL_685972\|2020-05 | .................... | .................... | ..................... | .................... | .................... | .................... |
| V | 3645 | hCoV-19/Japan/PG-2368/2020\|EPI_ISL_685973\|2020-05 | .................... | .................... | ..................... | .................... | .................... | .................... |
| V | 3646 | hCoV-19/Japan/PG-2369/2020\|EPI_ISL_685974\|2020-05 | .................... | .................... | ..................... | .................... | .................... | .................... |
| V | 3647 | hCoV-19/Japan/PG-2370/2020\|EPI_ISL_685975\|2020-05 | .................... | .................... | ..................... | .................... | .................... | .................... |
| V | 3648 | hCoV-19/Japan/PG-2371/2020\|EPI_ISL_685976\|2020-05 | .................... | .................... | ..................... | .................... | .................... | .................... |
| V | 3649 | hCoV-19/Japan/PG-2372/2020\|EPI_ISL_685977\|2020-05 | .................... | .................... | ..................... | .................... | .................... | .................... |
| V | 3650 | hCoV-19/Japan/PG-2373/2020\|EPI_ISL_685978\|2020-05 | .................... | .................... | ..................... | .................... | .................... | .................... |
| V | 3651 | hCoV-19/Japan/PG-2374/2020\|EPI_ISL_685979\|2020-05 | .................... | .................... | ..................... | .................... | .................... | .................... |
| V | 3652 | hCoV-19/Japan/PG-2375/2020\|EPI_ISL_685980\|2020-05 | .................... | .................... | ..................... | .................... | .................... | .................... |
| V | 3653 | hCoV-19/Japan/PG-2376/2020\|EPI_ISL_685981\|2020-05 | .................... | .................... | ..................... | .................... | .................... | .................... |
| V | 3655 | hCoV-19/Japan/PG-2378/2020\|EPI_ISL_685983\|2020-05 | .................... | .................... | ..................... | .................... | .................... | .................... |
| V | 3656 | hCoV-19/Japan/PG-2379/2020\|EPI_ISL_685984\|2020-05 | .................... | .................... | ..................... | .................... | .................... | .................... |
| V | 3657 | hCoV-19/Japan/PG-2380/2020\|EPI_ISL_685985\|2020-05 | .................... | .................... | ..................... | .................... | .................... | .................... |
| V | 3658 | hCoV-19/Japan/PG-2381/2020\|EPI_ISL_685986\|2020-05 | .................... | .................... | ..................... | .................... | .................... | .................... |
| V | 3659 | hCoV-19/Japan/PG-2382/2020\|EPI_ISL_685987\|2020-04 | .................... | .................... | ..................... | .................... | .................... | .................... |
| V | 3660 | hCoV-19/Japan/PG-2383/2020\|EPI_ISL_685988\|2020-03 | .................... | .................... | ..................... | .................... | .................... | .................... |
| V | 3661 | hCoV-19/Japan/PG-2384/2020\|EPI_ISL_685989\|2020-03 | .................... | .................... | ..................... | .................... | .................... | .................... |
| V | 3662 | hCoV-19/Japan/PG-2385/2020\|EPI_ISL_685990\|2020-03 | .................... | .................... | ..................... | .................... | .................... | .................... |
| V | 3663 | hCoV-19/Japan/PG-2386/2020\|EPI_ISL_685991\|2020-03 | .................... | .................... | ..................... | .................... | .................... | .................... |

| | | | | | | | | |
|---|---|---|---|---|---|---|---|---|
| V | 3664 | hCoV-19/Japan/PG-2387/2020\|EPI_ISL_685992\|2020-04 | .................... | .................... | ..................... | .................... | .................... | .................... |
| V | 3665 | hCoV-19/Japan/PG-2388/2020\|EPI_ISL_685993\|2020-04 | .................... | .................... | ..................... | .................... | .................... | .................... |
| V | 3666 | hCoV-19/Japan/PG-2389/2020\|EPI_ISL_685994\|2020-04 | .................... | .................... | ..................... | .................... | .................... | .................... |
| V | 3667 | hCoV-19/Japan/PG-2390/2020\|EPI_ISL_685995\|2020-04 | .................... | .................... | ..................... | .................... | .................... | .................... |
| V | 3668 | hCoV-19/Japan/PG-2391/2020\|EPI_ISL_685996\|2020-04 | .................... | .................... | ..................... | .................... | .................... | .................... |
| V | 3669 | hCoV-19/Japan/PG-2392/2020\|EPI_ISL_685997\|2020-04 | .................... | .................... | ..................... | .................... | .................... | .................... |
| V | 3670 | hCoV-19/Japan/PG-2393/2020\|EPI_ISL_685998\|2020-04 | .................... | .................... | ..................... | .................... | .................... | .................... |
| V | 3671 | hCoV-19/Japan/PG-2394/2020\|EPI_ISL_685999\|2020-04 | .................... | .................... | ..................... | .................... | .................... | .................... |
| V | 3672 | hCoV-19/Japan/PG-7134/2020\|EPI_ISL_688239\|2020-08 | .................... | .................... | ..................... | .................... | .................... | .................... |
| V | 3673 | hCoV-19/Japan/PG-7135/2020\|EPI_ISL_688240\|2020-08 | .................... | .................... | ..................... | .................... | .................... | .................... |
| V | 3674 | hCoV-19/Japan/PG-7136/2020\|EPI_ISL_688241\|2020-08 | .................... | .................... | ..................... | .................... | .................... | .................... |
| V | 3675 | hCoV-19/Japan/PG-7137/2020\|EPI_ISL_688242\|2020-08 | .................... | .................... | ..................... | .................... | .................... | .................... |
| V | 3676 | hCoV-19/Japan/PG-7138/2020\|EPI_ISL_688243\|2020-08 | .................... | .................... | ..................... | .................... | .................... | .................... |
| V | 3677 | hCoV-19/Japan/PG-7139/2020\|EPI_ISL_688244\|2020-08 | .................... | .................... | ..................... | .................... | .................... | .................... |
| V | 3678 | hCoV-19/Japan/PG-7140/2020\|EPI_ISL_688245\|2020-08 | .................... | .................... | ..................... | .................... | .................... | .................... |
| V | 3679 | hCoV-19/Japan/PG-7141/2020\|EPI_ISL_688246\|2020-08 | .................... | .................... | ..................... | .................... | .................... | .................... |
| V | 3680 | hCoV-19/Japan/PG-7142/2020\|EPI_ISL_688247\|2020-08 | .................... | .................... | ..................... | .................... | .................... | .................... |
| V | 3681 | hCoV-19/Japan/PG-7143/2020\|EPI_ISL_688248\|2020-08 | .................... | .................... | ..................... | .................... | .................... | .................... |
| V | 3682 | hCoV-19/Japan/PG-7144/2020\|EPI_ISL_688249\|2020-08 | .................... | .................... | ..................... | .................... | .................... | .................... |
| V | 3683 | hCoV-19/Japan/PG-7145/2020\|EPI_ISL_688250\|2020-08 | .................... | .................... | ..................... | .................... | .................... | .................... |
| V | 3684 | hCoV-19/Japan/PG-7146/2020\|EPI_ISL_688251\|2020-09 | .................... | .................... | ..................... | .................... | .................... | .................... |
| V | 3685 | hCoV-19/Japan/PG-7147/2020\|EPI_ISL_688252\|2020-09 | .................... | .................... | ..................... | .................... | .................... | .................... |
| V | 3686 | hCoV-19/Japan/PG-7148/2020\|EPI_ISL_688253\|2020-09 | .................... | .................... | ..................... | .................... | .................... | .................... |
| V | 3687 | hCoV-19/Japan/PG-7149/2020\|EPI_ISL_688254\|2020-09 | .................... | .................... | ..................... | .................... | .................... | .................... |
| V | 3688 | hCoV-19/Japan/PG-7150/2020\|EPI_ISL_688255\|2020-09 | .................... | .................... | ..................... | .................... | .................... | .................... |
| V | 3689 | hCoV-19/Japan/PG-7151/2020\|EPI_ISL_688256\|2020-09 | .................... | .................... | ..................... | .................... | .................... | .................... |
| V | 3690 | hCoV-19/Japan/PG-7152/2020\|EPI_ISL_688257\|2020-09 | .................... | .................... | ..................... | .................... | .................... | .................... |
| V | 3691 | hCoV-19/Japan/PG-7153/2020\|EPI_ISL_688258\|2020-09 | .................... | .................... | ..................... | .................... | .................... | .................... |
| V | 3692 | hCoV-19/Japan/PG-7154/2020\|EPI_ISL_688259\|2020-09 | .................... | .................... | ..................... | .................... | .................... | .................... |
| V | 3693 | hCoV-19/Japan/PG-7155/2020\|EPI_ISL_688260\|2020-09 | .................... | .................... | ..................... | .................... | .................... | .................... |
| V | 3694 | hCoV-19/Japan/PG-7156/2020\|EPI_ISL_688261\|2020-09 | .................... | .................... | ..................... | .................... | .................... | .................... |
| V | 3695 | hCoV-19/Japan/PG-7157/2020\|EPI_ISL_688262\|2020-09 | .................... | .................... | ..................... | .................... | .................... | .................... |
| V | 3696 | hCoV-19/Japan/PG-7158/2020\|EPI_ISL_688263\|2020-09 | .................... | .................... | ..................... | .................... | .................... | .................... |
| V | 3697 | hCoV-19/Japan/PG-7159/2020\|EPI_ISL_688264\|2020-09 | .................... | .................... | ..................... | .................... | .................... | .................... |
| V | 3698 | hCoV-19/Japan/PG-7160/2020\|EPI_ISL_688265\|2020-09 | .................... | .................... | ..................... | .................... | .................... | .................... |
| V | 3699 | hCoV-19/Japan/PG-7161/2020\|EPI_ISL_688266\|2020-09 | .................... | .................... | ..................... | .................... | .................... | .................... |
| V | 3700 | hCoV-19/Japan/PG-7162/2020\|EPI_ISL_688267\|2020-09 | .................... | .................... | ..................... | .................... | .................... | .................... |
| V | 3701 | hCoV-19/Japan/PG-7163/2020\|EPI_ISL_688268\|2020-09 | .................... | .................... | ..................... | .................... | .................... | .................... |
| V | 3702 | hCoV-19/Japan/PG-7164/2020\|EPI_ISL_688269\|2020-07 | .................... | .................... | ..................... | .................... | .................... | .................... |
| V | 3703 | hCoV-19/Japan/PG-7165/2020\|EPI_ISL_688270\|2020-07 | .................... | .................... | ..................... | .................... | .................... | .................... |
| V | 3704 | hCoV-19/Japan/PG-7166/2020\|EPI_ISL_688271\|2020-07 | .................... | .................... | ..................... | .................... | .................... | .................... |
| V | 3705 | hCoV-19/Japan/PG-7167/2020\|EPI_ISL_688272\|2020-07 | .................... | .................... | ..................... | .................... | .................... | .................... |
| V | 3706 | hCoV-19/Japan/PG-7168/2020\|EPI_ISL_688273\|2020-07 | .................... | .................... | ..................... | .................... | .................... | .................... |
| V | 3707 | hCoV-19/Japan/PG-7169/2020\|EPI_ISL_688274\|2020-07 | .................... | .................... | ..................... | .................... | .................... | .................... |
| V | 3708 | hCoV-19/Japan/PG-7170/2020\|EPI_ISL_688275\|2020-07 | .................... | .................... | ..................... | .................... | .................... | .................... |
| V | 3709 | hCoV-19/Japan/PG-7171/2020\|EPI_ISL_688276\|2020-07 | .................... | .................... | ..................... | .................... | .................... | .................... |
| V | 3710 | hCoV-19/Japan/PG-7172/2020\|EPI_ISL_688277\|2020-07 | .................... | .................... | ..................... | .................... | .................... | .................... |
| V | 3711 | hCoV-19/Japan/PG-7173/2020\|EPI_ISL_688278\|2020-07 | .................... | .................... | ..................... | .................... | .................... | .................... |
| V | 3712 | hCoV-19/Japan/PG-7174/2020\|EPI_ISL_688279\|2020-07 | .................... | .................... | ..................... | .................... | .................... | .................... |
| V | 3713 | hCoV-19/Japan/PG-6465/2020\|EPI_ISL_689570\|2020-08 | .................... | .................... | ..................... | .................... | .................... | .................... |
| V | 3714 | hCoV-19/Japan/PG-6466/2020\|EPI_ISL_689571\|2020-08 | .................... | .................... | ..................... | .................... | .................... | .................... |
| V | 3715 | hCoV-19/Japan/PG-6467/2020\|EPI_ISL_689572\|2020-08 | .................... | .................... | ..................... | .................... | .................... | .................... |
| V | 3716 | hCoV-19/Japan/PG-6468/2020\|EPI_ISL_689573\|2020-08 | .................... | .................... | ..................... | .................... | .................... | .................... |
| V | 3717 | hCoV-19/Japan/PG-6469/2020\|EPI_ISL_689574\|2020-08 | .................... | .................... | ..................... | .................... | .................... | .................... |
| V | 3718 | hCoV-19/Japan/PG-6470/2020\|EPI_ISL_689575\|2020-08 | .................... | .................... | ..................... | .................... | .................... | .................... |
| V | 3719 | hCoV-19/Japan/PG-6471/2020\|EPI_ISL_689576\|2020-08 | .................... | .................... | ..................... | .................... | .................... | .................... |
| V | 3720 | hCoV-19/Japan/PG-6472/2020\|EPI_ISL_689577\|2020-08 | .................... | .................... | ..................... | .................... | .................... | .................... |
| V | 3721 | hCoV-19/Japan/PG-6473/2020\|EPI_ISL_689578\|2020-08 | .................... | .................... | ..................... | .................... | .................... | .................... |
| V | 3722 | hCoV-19/Japan/PG-6474/2020\|EPI_ISL_689579\|2020-08 | .................... | .................... | ..................... | .................... | .................... | .................... |
| V | 3723 | hCoV-19/Japan/PG-6475/2020\|EPI_ISL_689580\|2020-08 | .................... | .................... | ..................... | .................... | .................... | .................... |
| V | 3724 | hCoV-19/Japan/PG-6476/2020\|EPI_ISL_689581\|2020-08 | .................... | .................... | ..................... | .................... | .................... | .................... |
| V | 3725 | hCoV-19/Japan/PG-6477/2020\|EPI_ISL_689582\|2020-08 | .................... | .................... | ..................... | .................... | .................... | .................... |
| V | 3726 | hCoV-19/Japan/PG-6478/2020\|EPI_ISL_689583\|2020-08 | .................... | .................... | ..................... | .................... | .................... | .................... |
| V | 3727 | hCoV-19/Japan/PG-6479/2020\|EPI_ISL_689584\|2020-08 | .................... | .................... | ..................... | .................... | .................... | .................... |
| V | 3728 | hCoV-19/Japan/PG-6480/2020\|EPI_ISL_689585\|2020-08 | .................... | .................... | ..................... | .................... | .................... | .................... |
| V | 3729 | hCoV-19/Japan/PG-6481/2020\|EPI_ISL_689586\|2020-08 | .................... | .................... | ..................... | .................... | .................... | .................... |
| V | 3730 | hCoV-19/Japan/PG-6482/2020\|EPI_ISL_689587\|2020-08 | .................... | .................... | ..................... | .................... | .................... | .................... |
| V | 3731 | hCoV-19/Japan/PG-6484/2020\|EPI_ISL_689588\|2020-08 | .................... | .................... | ..................... | .................... | .................... | .................... |
| V | 3732 | hCoV-19/Japan/PG-6485/2020\|EPI_ISL_689589\|2020-08 | .................... | .................... | ..................... | .................... | .................... | .................... |
| V | 3733 | hCoV-19/Japan/PG-6486/2020\|EPI_ISL_689590\|2020-08 | .................... | .................... | ..................... | .................... | .................... | .................... |
| V | 3734 | hCoV-19/Japan/PG-6489/2020\|EPI_ISL_689591\|2020-08 | .................... | .................... | ..................... | .................... | .................... | .................... |
| V | 3735 | hCoV-19/Japan/PG-6490/2020\|EPI_ISL_689592\|2020-09 | .................... | .................... | ..................... | .................... | .................... | .................... |
| V | 3736 | hCoV-19/Japan/PG-6491/2020\|EPI_ISL_689593\|2020-09 | .................... | .................... | ..................... | .................... | .................... | .................... |
| V | 3737 | hCoV-19/Japan/PG-6492/2020\|EPI_ISL_689594\|2020-09 | .................... | .................... | ..................... | .................... | .................... | .................... |
| V | 3738 | hCoV-19/Japan/PG-6493/2020\|EPI_ISL_689595\|2020-09 | .................... | .................... | ..................... | .................... | .................... | .................... |
| V | 3739 | hCoV-19/Japan/PG-6494/2020\|EPI_ISL_689596\|2020-09 | .................... | .................... | ..................... | .................... | .................... | .................... |
| V | 3740 | hCoV-19/Japan/PG-6495/2020\|EPI_ISL_689597\|2020-09 | .................... | .................... | ..................... | .................... | .................... | .................... |
| V | 3741 | hCoV-19/Japan/PG-6496/2020\|EPI_ISL_689598\|2020-09 | .................... | .................... | ..................... | .................... | .................... | .................... |
| V | 3742 | hCoV-19/Japan/PG-6497/2020\|EPI_ISL_689599\|2020-09 | .................... | .................... | ..................... | .................... | .................... | .................... |
| V | 3743 | hCoV-19/Japan/PG-5407/2020\|EPI_ISL_690510\|2020-08 | .................... | .................... | ..................... | .................... | .................... | .................... |

| | | | | | | | | |
|---|---|---|---|---|---|---|---|---|
| V | 3744 | hCoV-19/Japan/PG-5408/2020\|EPI_ISL_690511\|2020-08 | .................... | .................... | ..................... | .................... | .................... | .................... |
| V | 3745 | hCoV-19/Japan/PG-5409/2020\|EPI_ISL_690512\|2020-08 | .................... | .................... | ..................... | .................... | .................... | .................... |
| V | 3747 | hCoV-19/Japan/PG-5412/2020\|EPI_ISL_690515\|2020-08 | .................... | .................... | ..................... | .................... | .................... | .................... |
| V | 3749 | hCoV-19/Japan/PG-5414/2020\|EPI_ISL_690517\|2020-08 | .................... | .................... | ..................... | .................... | .................... | .................... |
| V | 3750 | hCoV-19/Japan/PG-5415/2020\|EPI_ISL_690518\|2020-08 | .................... | .................... | ..................... | .................... | .................... | .................... |
| V | 3751 | hCoV-19/Japan/PG-5416/2020\|EPI_ISL_690519\|2020-08 | .................... | .................... | ..................... | .................... | .................... | .................... |
| V | 3752 | hCoV-19/Japan/PG-5417/2020\|EPI_ISL_690520\|2020-08 | .................... | .................... | ..................... | .................... | .................... | .................... |
| V | 3753 | hCoV-19/Japan/PG-5418/2020\|EPI_ISL_690521\|2020-08 | .................... | .................... | ..................... | .................... | .................... | .................... |
| V | 3754 | hCoV-19/Japan/PG-5419/2020\|EPI_ISL_690522\|2020-08 | .................... | .................... | ..................... | .................... | .................... | .................... |
| V | 3755 | hCoV-19/Japan/PG-5420/2020\|EPI_ISL_690523\|2020-08 | .................... | .................... | ..................... | .................... | .................... | .................... |
| V | 3756 | hCoV-19/Japan/PG-5421/2020\|EPI_ISL_690524\|2020-08 | .................... | .................... | ..................... | .................... | .................... | .................... |
| V | 3757 | hCoV-19/Japan/PG-5422/2020\|EPI_ISL_690525\|2020-08 | .................... | .................... | ..................... | .................... | .................... | .................... |
| V | 3758 | hCoV-19/Japan/PG-5423/2020\|EPI_ISL_690526\|2020-08 | .................... | .................... | ..................... | .................... | .................... | .................... |
| V | 3759 | hCoV-19/Japan/PG-5424/2020\|EPI_ISL_690527\|2020-07 | .................... | .................... | ..................... | .................... | .................... | .................... |
| V | 3760 | hCoV-19/Japan/PG-5425/2020\|EPI_ISL_690528\|2020-07 | .................... | .................... | ..................... | .................... | .................... | .................... |
| V | 3761 | hCoV-19/Japan/PG-5426/2020\|EPI_ISL_690529\|2020-07 | .................... | .................... | ..................... | .................... | .................... | .................... |
| V | 3762 | hCoV-19/Japan/PG-5427/2020\|EPI_ISL_690530\|2020-08 | .................... | .................... | ..................... | .................... | .................... | .................... |
| V | 3763 | hCoV-19/Japan/PG-5428/2020\|EPI_ISL_690531\|2020-07 | .................... | .................... | ..................... | .................... | .................... | .................... |
| V | 3764 | hCoV-19/Japan/PG-5429/2020\|EPI_ISL_690532\|2020-07 | .................... | .................... | ..................... | .................... | .................... | .................... |
| V | 3765 | hCoV-19/Japan/PG-5430/2020\|EPI_ISL_690533\|2020-07 | .................... | .................... | ..................... | .................... | .................... | .................... |
| V | 3766 | hCoV-19/Japan/PG-5431/2020\|EPI_ISL_690534\|2020-07 | .................... | .................... | ..................... | .................... | .................... | .................... |
| V | 3767 | hCoV-19/Japan/PG-5432/2020\|EPI_ISL_690535\|2020-08 | .................... | .................... | ..................... | .................... | .................... | .................... |
| V | 3768 | hCoV-19/Japan/PG-5433/2020\|EPI_ISL_690536\|2020-08 | .................... | .................... | ..................... | .................... | .................... | .................... |
| V | 3769 | hCoV-19/Japan/PG-5434/2020\|EPI_ISL_690537\|2020-08 | .................... | .................... | ..................... | .................... | .................... | .................... |
| V | 3770 | hCoV-19/Japan/PG-5435/2020\|EPI_ISL_690538\|2020-08 | .................... | .................... | ..................... | .................... | .................... | .................... |
| V | 3771 | hCoV-19/Japan/PG-5436/2020\|EPI_ISL_690539\|2020-08 | .................... | .................... | ..................... | .................... | .................... | .................... |
| V | 3772 | hCoV-19/Japan/PG-5437/2020\|EPI_ISL_690540\|2020-08 | .................... | .................... | ..................... | .................... | .................... | .................... |
| V | 3773 | hCoV-19/Japan/PG-5438/2020\|EPI_ISL_690541\|2020-08 | .................... | .................... | ..................... | .................... | .................... | .................... |
| V | 3774 | hCoV-19/Japan/PG-5439/2020\|EPI_ISL_690542\|2020-08 | .................... | .................... | ..................... | .................... | .................... | .................... |
| V | 3775 | hCoV-19/Japan/PG-5440/2020\|EPI_ISL_690543\|2020-08 | .................... | .................... | ..................... | .................... | .................... | .................... |
| V | 3776 | hCoV-19/Japan/PG-5441/2020\|EPI_ISL_690544\|2020-08 | .................... | .................... | ..................... | .................... | .................... | .................... |
| V | 3777 | hCoV-19/Japan/PG-5442/2020\|EPI_ISL_690545\|2020-08 | .................... | .................... | ..................... | .................... | .................... | .................... |
| V | 3778 | hCoV-19/Japan/PG-5443/2020\|EPI_ISL_690546\|2020-08 | .................... | .................... | ..................... | .................... | .................... | .................... |
| V | 3779 | hCoV-19/Japan/PG-5444/2020\|EPI_ISL_690547\|2020-08 | .................... | .................... | ..................... | .................... | .................... | .................... |
| V | 3780 | hCoV-19/Japan/PG-5445/2020\|EPI_ISL_690548\|2020-08 | .................... | .................... | ..................... | .................... | .................... | .................... |
| V | 3781 | hCoV-19/Japan/PG-5446/2020\|EPI_ISL_690549\|2020-08 | .................... | .................... | ..................... | .................... | .................... | .................... |
| V | 3782 | hCoV-19/Japan/PG-3584/2020\|EPI_ISL_691840\|2020-06 | .................... | .................... | ..................... | .................... | .................... | .................... |
| V | 3783 | hCoV-19/Japan/PG-3585/2020\|EPI_ISL_691841\|2020-06 | .................... | .................... | ..................... | .................... | .................... | .................... |
| V | 3784 | hCoV-19/Japan/PG-3588/2020\|EPI_ISL_691842\|2020-07 | .................... | .................... | ..................... | .................... | .................... | .................... |
| V | 3785 | hCoV-19/Japan/PG-3589/2020\|EPI_ISL_691843\|2020-07 | .................... | .................... | ..................... | .................... | .................... | .................... |
| V | 3786 | hCoV-19/Japan/PG-3590/2020\|EPI_ISL_691844\|2020-07 | .................... | .................... | ..................... | .................... | .................... | .................... |
| V | 3787 | hCoV-19/Japan/PG-3591/2020\|EPI_ISL_691845\|2020-07 | .................... | .................... | ..................... | .................... | .................... | .................... |
| V | 3788 | hCoV-19/Japan/PG-3594/2020\|EPI_ISL_691848\|2020-07 | .................... | .................... | ..................... | .................... | .................... | .................... |
| V | 3789 | hCoV-19/Japan/PG-3595/2020\|EPI_ISL_691849\|2020-07 | .................... | .................... | ..................... | .................... | .................... | .................... |
| V | 3790 | hCoV-19/Japan/PG-3596/2020\|EPI_ISL_691850\|2020-07 | .................... | .................... | ..................... | .................... | .................... | .................... |
| V | 3791 | hCoV-19/Japan/PG-3597/2020\|EPI_ISL_691851\|2020-07 | .................... | .................... | ..................... | .................... | .................... | .................... |
| V | 3792 | hCoV-19/Japan/PG-3598/2020\|EPI_ISL_691852\|2020-07 | .................... | .................... | ..................... | .................... | .................... | .................... |
| V | 3793 | hCoV-19/Japan/PG-3599/2020\|EPI_ISL_691853\|2020-07 | .................... | .................... | ..................... | .................... | .................... | .................... |
| V | 3794 | hCoV-19/Japan/PG-3600/2020\|EPI_ISL_691854\|2020-07 | .................... | .................... | ..................... | .................... | .................... | .................... |
| V | 3795 | hCoV-19/Japan/PG-3601/2020\|EPI_ISL_691855\|2020-07 | .................... | .................... | ..................... | .................... | .................... | .................... |
| V | 3796 | hCoV-19/Japan/PG-3602/2020\|EPI_ISL_691856\|2020-07 | .................... | .................... | ..................... | .................... | .................... | .................... |
| V | 3797 | hCoV-19/Japan/PG-3603/2020\|EPI_ISL_691857\|2020-07 | .................... | .................... | ..................... | .................... | .................... | .................... |
| V | 3798 | hCoV-19/Japan/PG-3604/2020\|EPI_ISL_691858\|2020-07 | .................... | .................... | ..................... | .................... | .................... | .................... |
| V | 3799 | hCoV-19/Japan/PG-3605/2020\|EPI_ISL_691859\|2020-07 | .................... | .................... | ..................... | .................... | .................... | .................... |
| V | 3800 | hCoV-19/Japan/PG-3606/2020\|EPI_ISL_691860\|2020-07 | .................... | .................... | ..................... | .................... | .................... | .................... |
| V | 3801 | hCoV-19/Japan/PG-3607/2020\|EPI_ISL_691861\|2020-05 | .................... | .................... | ..................... | .................... | .................... | .................... |
| V | 3802 | hCoV-19/Japan/PG-3608/2020\|EPI_ISL_691862\|2020-06 | .................... | .................... | ..................... | .................... | .................... | .................... |
| V | 3803 | hCoV-19/Japan/PG-3609/2020\|EPI_ISL_691863\|2020-06 | .................... | .................... | ..................... | .................... | .................... | .................... |
| V | 3804 | hCoV-19/Japan/PG-3611/2020\|EPI_ISL_691864\|2020-07 | .................... | .................... | ..................... | .................... | .................... | .................... |
| V | 3805 | hCoV-19/Japan/PG-3612/2020\|EPI_ISL_691865\|2020-07 | .................... | .................... | ..................... | .................... | .................... | .................... |
| V | 3806 | hCoV-19/Japan/PG-3613/2020\|EPI_ISL_691866\|2020-07 | .................... | .................... | ..................... | .................... | .................... | .................... |
| V | 3807 | hCoV-19/Japan/PG-3614/2020\|EPI_ISL_691867\|2020-07 | .................... | .................... | ..................... | .................... | .................... | .................... |
| V | 3808 | hCoV-19/Japan/PG-3615/2020\|EPI_ISL_691868\|2020-07 | .................... | .................... | ..................... | .................... | .................... | .................... |
| V | 3809 | hCoV-19/Japan/PG-3616/2020\|EPI_ISL_691869\|2020-07 | .................... | .................... | ..................... | .................... | .................... | .................... |
| V | 3810 | hCoV-19/Japan/PG-3617/2020\|EPI_ISL_691870\|2020-07 | .................... | .................... | ..................... | .................... | .................... | .................... |
| V | 3811 | hCoV-19/Japan/PG-3618/2020\|EPI_ISL_691871\|2020-07 | .................... | .................... | ..................... | .................... | .................... | .................... |
| V | 3812 | hCoV-19/Japan/PG-3619/2020\|EPI_ISL_691872\|2020-04 | .................... | .................... | ..................... | .................... | .................... | .................... |
| V | 3813 | hCoV-19/Japan/PG-3620/2020\|EPI_ISL_691873\|2020-04 | .................... | .................... | ..................... | .................... | .................... | .................... |
| V | 3814 | hCoV-19/Japan/PG-3621/2020\|EPI_ISL_691874\|2020-04 | .................... | .................... | ..................... | .................... | .................... | .................... |
| V | 3815 | hCoV-19/Japan/PG-3622/2020\|EPI_ISL_691875\|2020-04 | .................... | .................... | ..................... | .................... | .................... | .................... |
| V | 3816 | hCoV-19/Japan/PG-3623/2020\|EPI_ISL_691876\|2020-04 | .................... | .................... | ..................... | .................... | .................... | .................... |
| V | 3817 | hCoV-19/Japan/PG-3624/2020\|EPI_ISL_691877\|2020-04 | .................... | .................... | ..................... | .................... | .................... | .................... |
| V | 3818 | hCoV-19/Japan/PG-3625/2020\|EPI_ISL_691878\|2020-04 | .................... | .................... | ..................... | .................... | .................... | .................... |
| V | 3819 | hCoV-19/Japan/PG-3626/2020\|EPI_ISL_691879\|2020-04 | .................... | .................... | ..................... | .................... | .................... | .................... |
| V | 3820 | hCoV-19/Switzerland/ZH-ETHZ-370518/2020\|EPI_ISL_69 | .................... | .................... | ..................... | .................... | .................... | .................... |
| V | 3821 | hCoV-19/Switzerland/ZH-ETHZ-370519/2020\|EPI_ISL_69 | .................... | .................... | ..................... | .................... | .................... | .................... |
| V | 3822 | hCoV-19/Switzerland/BE-ETHZ-370520/2020\|EPI_ISL_69 | .................... | ...............G.... | ..................... | .................... | .................... | .................... |
| V | 3823 | hCoV-19/Switzerland/BS-ETHZ-370521/2020\|EPI_ISL_69 | .................... | .................... | ..................... | .................... | .................... | .................... |
| V | 3824 | hCoV-19/Switzerland/AG-ETHZ-370522/2020\|EPI_ISL_69 | .................... | ......A............. | ..................... | .................... | .................... | .................... |
| V | 3825 | hCoV-19/Switzerland/AG-ETHZ-370524/2020\|EPI_ISL_69 | .................... | .................... | ..................... | .................... | .................... | .................... |

V 3826 hCoV-19/Switzerland/BE-ETHZ-370525/2020|EPI_ISL_69 .................... .................... .................... .................... .................... ....................
V 3827 hCoV-19/Switzerland/ZH-ETHZ-370526/2020|EPI_ISL_69 .................... .................... .................... .................... .................... ....................
V 3828 hCoV-19/Switzerland/AG-ETHZ-370527/2020|EPI_ISL_69 .................... .................... .................... .................... .................... ....................
V 3829 hCoV-19/USA/AZ-TG307287/2020|EPI_ISL_694009|2020-0 .................... .................... .................... .................... .................... ....................
V 3830 hCoV-19/USA/AZ-TG310521/2020|EPI_ISL_694010|2020-0 .................... .................... .................... .................... .................... ....................
V 3831 hCoV-19/USA/AZ-TG310526/2020|EPI_ISL_694011|2020-0 .................... .................... .................... .................... .................... ....................
V 3832 hCoV-19/USA/AZ-TG312407/2020|EPI_ISL_694013|2020-0 .................... .................... .................... .................... .................... ....................
V 3833 hCoV-19/USA/AZ-TG312835/2020|EPI_ISL_694014|2020-0 .................... .................... .................... .................... .................... ....................
V 3834 hCoV-19/USA/AZ-TG319953/2020|EPI_ISL_694016|2020-0 .................... .................... .................... .................... .................... ....................
V 3835 hCoV-19/USA/AZ-TG320614/2020|EPI_ISL_694017|2020-0 .................... .................... .................... .................... .................... ....................
V 3836 hCoV-19/USA/AZ-TG324844/2020|EPI_ISL_694021|2020-0 .................... .................... .................... .................... .................... ....................
V 3837 hCoV-19/USA/AZ-TG329217/2020|EPI_ISL_694030|2020-0 .................... .................... .................... .................... .................... ....................
V 3838 hCoV-19/USA/AZ-TG329289/2020|EPI_ISL_694031|2020-0 .................... .................... .................... .................... .................... ....................
V 3839 hCoV-19/USA/AZ-TG333625/2020|EPI_ISL_694033|2020-0 .................... .................... .................... .................... .................... ....................
V 3840 hCoV-19/USA/AZ-TG357967/2020|EPI_ISL_694039|2020-0 .................... .................... .................... .................... .................... ....................
V 3841 hCoV-19/Japan/PG-0990/2020|EPI_ISL_684600|2020-04 .................... .................... .................... .................... .................... ....................
V 3842 hCoV-19/Japan/PG-0991/2020|EPI_ISL_684601|2020-04 .................... .................... .................... .................... .................... ....................
V 3843 hCoV-19/Japan/PG-0992/2020|EPI_ISL_684602|2020-04 .................... .................... .................... .................... .................... ....................
V 3844 hCoV-19/Japan/PG-0993/2020|EPI_ISL_684603|2020-04 .................... .................... .................... .................... .................... ....................
V 3845 hCoV-19/Japan/PG-0994/2020|EPI_ISL_684604|2020-04 .................... .................... .................... .................... .................... ....................
V 3846 hCoV-19/Japan/PG-0995/2020|EPI_ISL_684605|2020-04 .................... .................... .................... .................... .................... ....................
V 3847 hCoV-19/Japan/PG-0996/2020|EPI_ISL_684606|2020-04 .................... .................... .................... .................... .................... ....................
V 3848 hCoV-19/Japan/PG-0997/2020|EPI_ISL_684607|2020-04 .................... .................... .................... .................... .................... ....................
V 3849 hCoV-19/Japan/PG-0998/2020|EPI_ISL_684608|2020-04 .................... .................... .................... .................... .................... ....................
V 3850 hCoV-19/Japan/PG-0999/2020|EPI_ISL_684609|2020-04 .................... .................... .................... .................... .................... ....................
V 3851 hCoV-19/Japan/PG-1000/2020|EPI_ISL_684610|2020-04 .................... .................... .................... .................... .................... ....................
V 3852 hCoV-19/Japan/PG-1001/2020|EPI_ISL_684611|2020-04 .................... .................... .................... .................... .................... ....................
V 3853 hCoV-19/Japan/PG-1002/2020|EPI_ISL_684612|2020-04 .................... .................... .................... .................... .................... ....................
V 3854 hCoV-19/Japan/PG-1003/2020|EPI_ISL_684613|2020-04 .................... .................... .................... .................... .................... ....................
V 3855 hCoV-19/Japan/PG-1004/2020|EPI_ISL_684614|2020-04 .................... .................... .................... .................... .................... ....................
V 3856 hCoV-19/Japan/PG-1005/2020|EPI_ISL_684615|2020-02 .................... .................... .................... .................... .................... ....................
V 3857 hCoV-19/Japan/PG-1006/2020|EPI_ISL_684616|2020-02 .................... .................... .................... .................... .................... ....................
V 3858 hCoV-19/Japan/PG-1007/2020|EPI_ISL_684617|2020-03 .................... .................... .................... .................... .................... ....................
V 3859 hCoV-19/Japan/PG-1009/2020|EPI_ISL_684619|2020-03 .................... .................... .................... .................... .................... ....................
V 3860 hCoV-19/Japan/PG-1010/2020|EPI_ISL_684620|2020-03 .................... .................... .................... .................... .................... ....................
V 3861 hCoV-19/Japan/PG-1012/2020|EPI_ISL_684622|2020-03 .................... .................... .................... .................... .................... ....................
V 3862 hCoV-19/Japan/PG-1013/2020|EPI_ISL_684623|2020-03 .................... .................... .................... .................... .................... ....................
V 3863 hCoV-19/Japan/PG-1014/2020|EPI_ISL_684624|2020-03 .................... .................... .................... .................... .................... ....................
V 3864 hCoV-19/Japan/PG-1015/2020|EPI_ISL_684625|2020-03 .................... .................... .................... .................... .................... ....................
V 3865 hCoV-19/Japan/PG-1016/2020|EPI_ISL_684626|2020-03 .................... .................... .................... .................... .................... ....................
V 3866 hCoV-19/Japan/PG-1017/2020|EPI_ISL_684627|2020-03 .................... .................... .................... .................... .................... ....................
V 3867 hCoV-19/Japan/PG-1065/2020|EPI_ISL_684675|2020-04 .................... .................... .................... .................... .................... ....................
V 3868 hCoV-19/Japan/PG-1066/2020|EPI_ISL_684676|2020-04 .................... .................... .................... .................... .................... ....................
V 3869 hCoV-19/Japan/PG-1067/2020|EPI_ISL_684677|2020-04 .................... .................... .................... .................... .................... ....................
V 3870 hCoV-19/Japan/PG-1068/2020|EPI_ISL_684678|2020-04 .................... .................... .................... .................... .................... ....................
V 3871 hCoV-19/Japan/PG-1069/2020|EPI_ISL_684679|2020-04 .................... .................... .................... .................... .................... ....................
V 3872 hCoV-19/Japan/PG-1070/2020|EPI_ISL_684680|2020-04 .................... .................... .................... .................... .................... ....................
V 3873 hCoV-19/Japan/PG-1071/2020|EPI_ISL_684681|2020-04 .................... .................... .................... .................... .................... ....................
V 3874 hCoV-19/Japan/PG-1072/2020|EPI_ISL_684682|2020-04 .................... .................... .................... .................... .................... ....................
V 3875 hCoV-19/Japan/PG-1073/2020|EPI_ISL_684683|2020-04 .................... .................... .................... .................... .................... ....................
V 3876 hCoV-19/Japan/PG-1074/2020|EPI_ISL_684684|2020-04 .................... .................... .................... .................... .................... ....................
V 3877 hCoV-19/Japan/PG-1075/2020|EPI_ISL_684685|2020-04 .................... .................... .................... .................... .................... ....................
V 3878 hCoV-19/Japan/PG-1076/2020|EPI_ISL_684686|2020-04 .................... .................... .................... .................... .................... ....................
V 3879 hCoV-19/Japan/PG-1077/2020|EPI_ISL_684687|2020-04 .................... .................... .................... .................... .................... ....................
V 3880 hCoV-19/Japan/PG-1078/2020|EPI_ISL_684688|2020-04 .................... .................... .................... .................... .................... ....................
V 3881 hCoV-19/Japan/PG-1079/2020|EPI_ISL_684689|2020-04 .................... .................... .................... .................... .................... ....................
V 3882 hCoV-19/Japan/PG-1080/2020|EPI_ISL_684690|2020-04 .................... .................... .................... .................... .................... ....................
V 3883 hCoV-19/Japan/PG-1081/2020|EPI_ISL_684691|2020-04 .................... .................... .................... .................... .................... ....................
V 3884 hCoV-19/Japan/PG-1082/2020|EPI_ISL_684692|2020-04 .................... .................... .................... .................... .................... ....................
V 3885 hCoV-19/Japan/PG-1083/2020|EPI_ISL_684693|2020-03 .................... .................... .................... .................... .................... ....................
V 3886 hCoV-19/Japan/PG-1084/2020|EPI_ISL_684694|2020-03 .................... .................... .................... .................... .................... ....................
V 3887 hCoV-19/Japan/PG-1085/2020|EPI_ISL_684695|2020-04 .................... .................... .................... .................... .................... ....................
V 3888 hCoV-19/Japan/PG-1086/2020|EPI_ISL_684696|2020-04 .................... .................... .................... .................... .................... ....................
V 3889 hCoV-19/Japan/PG-1087/2020|EPI_ISL_684697|2020-04 .................... .................... .................... .................... .................... ....................
V 3890 hCoV-19/Japan/PG-1088/2020|EPI_ISL_684698|2020-04 .................... .................... .................... .................... .................... ....................
V 3891 hCoV-19/Japan/PG-1089/2020|EPI_ISL_684699|2020-04 .................... .................... .................... .................... .................... ....................
V 3892 hCoV-19/Japan/PG-2315/2020|EPI_ISL_685920|2020-04 .................... .................... .................... .................... .................... ....................
V 3893 hCoV-19/Japan/PG-2316/2020|EPI_ISL_685921|2020-04 .................... .................... .................... .................... .................... ....................
V 3894 hCoV-19/Japan/PG-2317/2020|EPI_ISL_685922|2020-05 .................... .................... .................... .................... .................... ....................
V 3895 hCoV-19/Japan/PG-2319/2020|EPI_ISL_685924|2020-05 .................... .................... .................... .................... .................... ....................
V 3896 hCoV-19/Japan/PG-2320/2020|EPI_ISL_685925|2020-04 .................... .................... .................... .................... .................... ....................
V 3897 hCoV-19/Japan/PG-2321/2020|EPI_ISL_685926|2020-04 .................... .................... .................... .................... .................... ....................
V 3898 hCoV-19/Japan/PG-2322/2020|EPI_ISL_685927|2020-04 .................... .................... .................... .................... .................... ....................
V 3899 hCoV-19/Japan/PG-2323/2020|EPI_ISL_685928|2020-05 .................... .................... .................... .................... .................... ....................
V 3900 hCoV-19/Japan/PG-2324/2020|EPI_ISL_685929|2020-04 .................... .................... .................... .................... .................... ....................
V 3901 hCoV-19/Japan/PG-2325/2020|EPI_ISL_685930|2020-03 .................... .................... .................... .................... .................... ....................
V 3902 hCoV-19/Japan/PG-2326/2020|EPI_ISL_685931|2020-04 .................... .................... .................... .................... .................... ....................
V 3903 hCoV-19/Japan/PG-2327/2020|EPI_ISL_685932|2020-04 .................... .................... .................... .................... .................... ....................
V 3904 hCoV-19/Japan/PG-2328/2020|EPI_ISL_685933|2020-04 .................... .................... .................... .................... .................... ....................
V 3905 hCoV-19/Japan/PG-2329/2020|EPI_ISL_685934|2020-04 .................... .................... .................... .................... .................... ....................

| | | | | | | | | |
|---|---|---|---|---|---|---|---|---|
| V | 3906 | hCoV-19/Japan/PG-2330/2020\|EPI_ISL_685935\|2020-04 | ···················· | ···················· | ····················· | ···················· | ···················· | ···················· |
| V | 3907 | hCoV-19/Japan/PG-2331/2020\|EPI_ISL_685936\|2020-04 | ···················· | ···················· | ····················· | ···················· | ···················· | ···················· |
| V | 3908 | hCoV-19/Japan/PG-2332/2020\|EPI_ISL_685937\|2020-05 | ···················· | ···················· | ····················· | ···················· | ···················· | ···················· |
| V | 3909 | hCoV-19/Japan/PG-2333/2020\|EPI_ISL_685938\|2020-05 | ···················· | ······A············· | ····················· | ···················· | ···················· | ···················· |
| V | 3910 | hCoV-19/Japan/PG-2334/2020\|EPI_ISL_685939\|2020-05 | ···················· | ···················· | ····················· | ···················· | ···················· | ···················· |
| V | 3911 | hCoV-19/Japan/PG-2335/2020\|EPI_ISL_685940\|2020-05 | ···················· | ···················· | ····················· | ···················· | ···················· | ···················· |
| V | 3912 | hCoV-19/Japan/PG-2336/2020\|EPI_ISL_685941\|2020-05 | ···················· | ···················· | ····················· | ···················· | ···················· | ···················· |
| V | 3913 | hCoV-19/Japan/PG-2337/2020\|EPI_ISL_685942\|2020-05 | ···················· | ···················· | ····················· | ···················· | ···················· | ···················· |
| V | 3914 | hCoV-19/Japan/PG-2338/2020\|EPI_ISL_685943\|2020-05 | ···················· | ···················· | ····················· | ···················· | ···················· | ···················· |
| V | 3915 | hCoV-19/Japan/PG-2339/2020\|EPI_ISL_685944\|2020-05 | ···················· | ···················· | ····················· | ···················· | ···················· | ···················· |
| V | 3916 | hCoV-19/Japan/PG-2340/2020\|EPI_ISL_685945\|2020-05 | ···················· | ···················· | ····················· | ···················· | ···················· | ···················· |
| V | 3917 | hCoV-19/Japan/PG-2341/2020\|EPI_ISL_685946\|2020-05 | ···················· | ···················· | ····················· | ···················· | ···················· | ···················· |
| V | 3918 | hCoV-19/Japan/PG-2342/2020\|EPI_ISL_685947\|2020-04 | ···················· | ···················· | ····················· | ···················· | ···················· | ···················· |
| V | 3919 | hCoV-19/Japan/PG-2343/2020\|EPI_ISL_685948\|2020-05 | ···················· | ······A············· | ····················· | ···················· | ···················· | ···················· |
| V | 3920 | hCoV-19/Japan/PG-2344/2020\|EPI_ISL_685949\|2020-05 | ···················· | ···················· | ····················· | ···················· | ···················· | ···················· |
| V | 3921 | hCoV-19/Japan/PG-2345/2020\|EPI_ISL_685950\|2020-05 | ···················· | ···················· | ····················· | ···················· | ···················· | ···················· |
| V | 3922 | hCoV-19/Japan/PG-2346/2020\|EPI_ISL_685951\|2020-05 | ···················· | ···················· | ····················· | ···················· | ···················· | ···················· |
| V | 3923 | hCoV-19/Japan/PG-2347/2020\|EPI_ISL_685952\|2020-05 | ···················· | ···················· | ····················· | ···················· | ···················· | ···················· |
| V | 3924 | hCoV-19/Japan/PG-2348/2020\|EPI_ISL_685953\|2020-04 | ···················· | ···················· | ····················· | ···················· | ···················· | ···················· |
| V | 3926 | hCoV-19/Japan/PG-2350/2020\|EPI_ISL_685955\|2020-04 | ···················· | ···················· | ····················· | ···················· | ···················· | ···················· |
| V | 3927 | hCoV-19/Japan/PG-2351/2020\|EPI_ISL_685956\|2020-05 | ···················· | ···················· | ····················· | ···················· | ···················· | ···················· |
| V | 3928 | hCoV-19/Japan/PG-2352/2020\|EPI_ISL_685957\|2020-04 | ···················· | ···················· | ····················· | ···················· | ···················· | ···················· |
| V | 3930 | hCoV-19/Japan/PG-6995/2020\|EPI_ISL_688100\|2020-08 | ···················· | ···················· | ····················· | ···················· | ···················· | ···················· |
| V | 3931 | hCoV-19/Japan/PG-6996/2020\|EPI_ISL_688101\|2020-09 | ···················· | ···················· | ····················· | ···················· | ···················· | ···················· |
| V | 3932 | hCoV-19/Japan/PG-6997/2020\|EPI_ISL_688102\|2020-09 | ···················· | ···················· | ····················· | ···················· | ···················· | ···················· |
| V | 3933 | hCoV-19/Japan/PG-6998/2020\|EPI_ISL_688103\|2020-09 | ···················· | ···················· | ····················· | ···················· | ···················· | ···················· |
| V | 3934 | hCoV-19/Japan/PG-6999/2020\|EPI_ISL_688104\|2020-09 | ···················· | ···················· | ····················· | ···················· | ···················· | ···················· |
| V | 3935 | hCoV-19/Japan/PG-7000/2020\|EPI_ISL_688105\|2020-09 | ···················· | ···················· | ····················· | ···················· | ···················· | ···················· |
| V | 3936 | hCoV-19/Japan/PG-7001/2020\|EPI_ISL_688106\|2020-09 | ···················· | ···················· | ····················· | ···················· | ···················· | ···················· |
| V | 3937 | hCoV-19/Japan/PG-7002/2020\|EPI_ISL_688107\|2020-09 | ···················· | ···················· | ····················· | ···················· | ···················· | ···················· |
| V | 3938 | hCoV-19/Japan/PG-7003/2020\|EPI_ISL_688108\|2020-09 | ···················· | ···················· | ····················· | ···················· | ···················· | ···················· |
| V | 3939 | hCoV-19/Japan/PG-7004/2020\|EPI_ISL_688109\|2020-08 | ···················· | ···················· | ····················· | ···················· | ···················· | ···················· |
| V | 3940 | hCoV-19/Japan/PG-7005/2020\|EPI_ISL_688110\|2020-08 | ···················· | ···················· | ····················· | ···················· | ···················· | ···················· |
| V | 3941 | hCoV-19/Japan/PG-7006/2020\|EPI_ISL_688111\|2020-08 | ···················· | ···················· | ····················· | ···················· | ···················· | ···················· |
| V | 3942 | hCoV-19/Japan/PG-7007/2020\|EPI_ISL_688112\|2020-08 | ···················· | ···················· | ····················· | ···················· | ···················· | ···················· |
| V | 3943 | hCoV-19/Japan/PG-7008/2020\|EPI_ISL_688113\|2020-08 | ···················· | ···················· | ····················· | ···················· | ···················· | ···················· |
| V | 3944 | hCoV-19/Japan/PG-7009/2020\|EPI_ISL_688114\|2020-08 | ···················· | ···················· | ····················· | ···················· | ···················· | ···················· |
| V | 3945 | hCoV-19/Japan/PG-7010/2020\|EPI_ISL_688115\|2020-08 | ···················· | ···················· | ····················· | ···················· | ···················· | ···················· |
| V | 3946 | hCoV-19/Japan/PG-7011/2020\|EPI_ISL_688116\|2020-08 | ···················· | ···················· | ····················· | ···················· | ···················· | ···················· |
| V | 3947 | hCoV-19/Japan/PG-7012/2020\|EPI_ISL_688117\|2020-08 | ···················· | ···················· | ····················· | ···················· | ···················· | ···················· |
| V | 3948 | hCoV-19/Japan/PG-6289/2020\|EPI_ISL_689400\|2020-07 | ···················· | ···················· | ····················· | ···················· | ···················· | ···················· |
| V | 3949 | hCoV-19/Japan/PG-6290/2020\|EPI_ISL_689401\|2020-07 | ···················· | ···················· | ····················· | ···················· | ···················· | ···················· |
| V | 3950 | hCoV-19/Japan/PG-6291/2020\|EPI_ISL_689402\|2020-07 | ···················· | ···················· | ····················· | ···················· | ···················· | ···················· |
| V | 3951 | hCoV-19/Japan/PG-6292/2020\|EPI_ISL_689403\|2020-07 | ···················· | ···················· | ····················· | ···················· | ···················· | ···················· |
| V | 3952 | hCoV-19/Japan/PG-6293/2020\|EPI_ISL_689404\|2020-07 | ···················· | ···················· | ····················· | ···················· | ···················· | ···················· |
| V | 3953 | hCoV-19/Japan/PG-6294/2020\|EPI_ISL_689405\|2020-07 | ···················· | ···················· | ····················· | ···················· | ···················· | ···················· |
| V | 3954 | hCoV-19/Japan/PG-6295/2020\|EPI_ISL_689406\|2020-07 | ···················· | ···················· | ····················· | ···················· | ···················· | ···················· |
| V | 3955 | hCoV-19/Japan/PG-6296/2020\|EPI_ISL_689407\|2020-07 | ···················· | ···················· | ····················· | ···················· | ···················· | ···················· |
| V | 3956 | hCoV-19/Japan/PG-6297/2020\|EPI_ISL_689408\|2020-07 | ···················· | ···················· | ····················· | ···················· | ···················· | ···················· |
| V | 3957 | hCoV-19/Japan/PG-6298/2020\|EPI_ISL_689409\|2020-08 | ···················· | ···················· | ····················· | ···················· | ···················· | ···················· |
| V | 3958 | hCoV-19/Japan/PG-6299/2020\|EPI_ISL_689410\|2020-08 | ···················· | ···················· | ····················· | ···················· | ···················· | ···················· |
| V | 3959 | hCoV-19/Japan/PG-6300/2020\|EPI_ISL_689411\|2020-08 | ···················· | ···················· | ····················· | ···················· | ···················· | ···················· |
| V | 3960 | hCoV-19/Japan/PG-6301/2020\|EPI_ISL_689412\|2020-08 | ···················· | ···················· | ····················· | ···················· | ···················· | ···················· |
| V | 3961 | hCoV-19/Japan/PG-6302/2020\|EPI_ISL_689413\|2020-08 | ···················· | ···················· | ····················· | ···················· | ···················· | ···················· |
| V | 3962 | hCoV-19/Japan/PG-6303/2020\|EPI_ISL_689414\|2020-08 | ···················· | ···················· | ····················· | ···················· | ···················· | ···················· |
| V | 3963 | hCoV-19/Japan/PG-6304/2020\|EPI_ISL_689415\|2020-08 | ···················· | ···················· | ····················· | ···················· | ···················· | ···················· |
| V | 3965 | hCoV-19/Japan/PG-6306/2020\|EPI_ISL_689417\|2020-08 | ···················· | ···················· | ····················· | ···················· | ···················· | ···················· |
| V | 3967 | hCoV-19/Japan/PG-6308/2020\|EPI_ISL_689419\|2020-08 | ···················· | ···················· | ····················· | ···················· | ···················· | ···················· |
| V | 3968 | hCoV-19/Japan/PG-6309/2020\|EPI_ISL_689420\|2020-08 | ···················· | ···················· | ····················· | ···················· | ···················· | ···················· |
| V | 3969 | hCoV-19/Japan/PG-6310/2020\|EPI_ISL_689421\|2020-08 | ···················· | ···················· | ····················· | ···················· | ···················· | ···················· |
| V | 3970 | hCoV-19/Japan/PG-6311/2020\|EPI_ISL_689422\|2020-08 | ···················· | ···················· | ····················· | ···················· | ···················· | ···················· |
| V | 3971 | hCoV-19/Japan/PG-6312/2020\|EPI_ISL_689423\|2020-08 | ···················· | ···················· | ····················· | ···················· | ···················· | ···················· |
| V | 3972 | hCoV-19/Japan/PG-6313/2020\|EPI_ISL_689424\|2020-08 | ···················· | ···················· | ····················· | ···················· | ···················· | ···················· |
| V | 3973 | hCoV-19/Japan/PG-6314/2020\|EPI_ISL_689425\|2020-07 | ···················· | ···················· | ····················· | ···················· | ···················· | ···················· |
| V | 3974 | hCoV-19/Japan/PG-6315/2020\|EPI_ISL_689426\|2020-08 | ···················· | ···················· | ····················· | ···················· | ···················· | ···················· |
| V | 3975 | hCoV-19/Japan/PG-6316/2020\|EPI_ISL_689427\|2020-08 | ···················· | ···················· | ····················· | ···················· | ···················· | ···················· |
| V | 3976 | hCoV-19/Japan/PG-6317/2020\|EPI_ISL_689428\|2020-08 | ···················· | ···················· | ····················· | ···················· | ···················· | ···················· |
| V | 3977 | hCoV-19/Japan/PG-6318/2020\|EPI_ISL_689429\|2020-08 | ···················· | ···················· | ····················· | ···················· | ···················· | ···················· |
| V | 3978 | hCoV-19/Japan/PG-6319/2020\|EPI_ISL_689430\|2020-08 | ···················· | ···················· | ····················· | ···················· | ···················· | ···················· |
| V | 3980 | hCoV-19/Japan/PG-6321/2020\|EPI_ISL_689432\|2020-08 | ···················· | ···················· | ····················· | ···················· | ···················· | ···················· |
| V | 3981 | hCoV-19/Japan/PG-6322/2020\|EPI_ISL_689433\|2020-08 | ···················· | ···················· | ····················· | ···················· | ···················· | ···················· |
| V | 3982 | hCoV-19/Japan/PG-6323/2020\|EPI_ISL_689434\|2020-08 | ···················· | ···················· | ····················· | ···················· | ···················· | ···················· |
| V | 3983 | hCoV-19/Japan/PG-6324/2020\|EPI_ISL_689435\|2020-07 | ···················· | ···················· | ····················· | ···················· | ···················· | ···················· |
| V | 3984 | hCoV-19/Japan/PG-6325/2020\|EPI_ISL_689436\|2020-07 | ···················· | ···················· | ····················· | ···················· | ···················· | ···················· |
| V | 3985 | hCoV-19/Japan/PG-6326/2020\|EPI_ISL_689437\|2020-07 | ···················· | ···················· | ····················· | ···················· | ···················· | ···················· |
| V | 3986 | hCoV-19/Japan/PG-6327/2020\|EPI_ISL_689438\|2020-07 | ···················· | ···················· | ····················· | ···················· | ···················· | ···················· |
| V | 3987 | hCoV-19/Japan/PG-6328/2020\|EPI_ISL_689439\|2020-07 | ···················· | ···················· | ····················· | ···················· | ···················· | ···················· |
| V | 3988 | hCoV-19/Japan/PG-6329/2020\|EPI_ISL_689440\|2020-07 | ···················· | ···················· | ····················· | ···················· | ···················· | ···················· |
| V | 3989 | hCoV-19/Japan/PG-6330/2020\|EPI_ISL_689441\|2020-07 | ···················· | ···················· | ····················· | ···················· | ···················· | ···················· |
| V | 3990 | hCoV-19/Japan/PG-6331/2020\|EPI_ISL_689442\|2020-07 | ···················· | ···················· | ····················· | ···················· | ···················· | ···················· |

| | | | | | | | | |
|---|---|---|---|---|---|---|---|---|
| V | 3991 | hCoV-19/Japan/PG-6332/2020\|EPI_ISL_689443\|2020-07 | ···················· | ···················· | ····················· | ···················· | ···················· | ···················· |
| V | 3992 | hCoV-19/Japan/PG-6333/2020\|EPI_ISL_689444\|2020-08 | ···················· | ···················· | ····················· | ···················· | ···················· | ···················· |
| V | 3993 | hCoV-19/Japan/PG-6334/2020\|EPI_ISL_689445\|2020-08 | ···················· | ···················· | ····················· | ···················· | ···················· | ···················· |
| V | 3994 | hCoV-19/Japan/PG-6335/2020\|EPI_ISL_689446\|2020-08 | ···················· | ···················· | ····················· | ···················· | ···················· | ···················· |
| V | 3995 | hCoV-19/Japan/PG-6336/2020\|EPI_ISL_689447\|2020-08 | ···················· | ···················· | ····················· | ···················· | ···················· | ···················· |
| V | 3996 | hCoV-19/Japan/PG-6337/2020\|EPI_ISL_689448\|2020-08 | ···················· | ···················· | ····················· | ···················· | ···················· | ···················· |
| V | 3997 | hCoV-19/Japan/PG-5327/2020\|EPI_ISL_690430\|2020-08 | ···················· | ···················· | ····················· | ···················· | ···················· | ···················· |
| V | 3998 | hCoV-19/Japan/PG-5328/2020\|EPI_ISL_690431\|2020-08 | ···················· | ···················· | ····················· | ···················· | ···················· | ···················· |
| V | 3999 | hCoV-19/Japan/PG-5329/2020\|EPI_ISL_690432\|2020-08 | ···················· | ···················· | ····················· | ···················· | ···················· | ···················· |
| V | 4000 | hCoV-19/Japan/PG-5330/2020\|EPI_ISL_690433\|2020-08 | ···················· | ···················· | ····················· | ···················· | ···················· | ···················· |
| V | 4001 | hCoV-19/Japan/PG-5331/2020\|EPI_ISL_690434\|2020-08 | ···················· | ···················· | ····················· | ···················· | ···················· | ···················· |
| V | 4002 | hCoV-19/Japan/PG-5332/2020\|EPI_ISL_690435\|2020-08 | ···················· | ···················· | ····················· | ···················· | ···················· | ···················· |
| V | 4003 | hCoV-19/Japan/PG-5333/2020\|EPI_ISL_690436\|2020-08 | ···················· | ···················· | ····················· | ···················· | ···················· | ···················· |
| V | 4004 | hCoV-19/Japan/PG-5334/2020\|EPI_ISL_690437\|2020-08 | ···················· | ···················· | ····················· | ···················· | ···················· | ···················· |
| V | 4005 | hCoV-19/Japan/PG-5335/2020\|EPI_ISL_690438\|2020-08 | ···················· | ···················· | ····················· | ···················· | ···················· | ···················· |
| V | 4006 | hCoV-19/Japan/PG-5336/2020\|EPI_ISL_690439\|2020-08 | ···················· | ···················· | ····················· | ···················· | ···················· | ···················· |
| V | 4007 | hCoV-19/Japan/PG-5337/2020\|EPI_ISL_690440\|2020-08 | ···················· | ···················· | ····················· | ···················· | ···················· | ···················· |
| V | 4008 | hCoV-19/Japan/PG-5338/2020\|EPI_ISL_690441\|2020-08 | ···················· | ···················· | ····················· | ···················· | ···················· | ···················· |
| V | 4009 | hCoV-19/Japan/PG-5339/2020\|EPI_ISL_690442\|2020-08 | ···················· | ···················· | ····················· | ···················· | ···················· | ···················· |
| V | 4010 | hCoV-19/Japan/PG-5340/2020\|EPI_ISL_690443\|2020-08 | ···················· | ···················· | ····················· | ···················· | ···················· | ···················· |
| V | 4011 | hCoV-19/Japan/PG-5341/2020\|EPI_ISL_690444\|2020-08 | ···················· | ···················· | ····················· | ···················· | ···················· | ···················· |
| V | 4012 | hCoV-19/Japan/PG-5342/2020\|EPI_ISL_690445\|2020-08 | ···················· | ···················· | ····················· | ···················· | ···················· | ···················· |
| V | 4013 | hCoV-19/Japan/PG-5343/2020\|EPI_ISL_690446\|2020-08 | ···················· | ···················· | ····················· | ···················· | ···················· | ···················· |
| V | 4014 | hCoV-19/Japan/PG-5344/2020\|EPI_ISL_690447\|2020-08 | ···················· | ···················· | ····················· | ···················· | ···················· | ···················· |
| V | 4015 | hCoV-19/Japan/PG-5345/2020\|EPI_ISL_690448\|2020-08 | ···················· | ···················· | ····················· | ···················· | ···················· | ···················· |
| V | 4016 | hCoV-19/Japan/PG-5346/2020\|EPI_ISL_690449\|2020-08 | ···················· | ···················· | ····················· | ···················· | ···················· | ···················· |
| V | 4017 | hCoV-19/Japan/PG-5347/2020\|EPI_ISL_690450\|2020-08 | ···················· | ···················· | ····················· | ···················· | ···················· | ···················· |
| V | 4018 | hCoV-19/Japan/PG-5348/2020\|EPI_ISL_690451\|2020-08 | ···················· | ···················· | ····················· | ···················· | ···················· | ···················· |
| V | 4019 | hCoV-19/Japan/PG-5349/2020\|EPI_ISL_690452\|2020-08 | ···················· | ···················· | ····················· | ···················· | ···················· | ···················· |
| V | 4020 | hCoV-19/Japan/PG-5350/2020\|EPI_ISL_690453\|2020-08 | ···················· | ···················· | ····················· | ···················· | ···················· | ···················· |
| V | 4021 | hCoV-19/Japan/PG-5351/2020\|EPI_ISL_690454\|2020-08 | ···················· | ···················· | ····················· | ···················· | ···················· | ···················· |
| V | 4022 | hCoV-19/Japan/PG-5352/2020\|EPI_ISL_690455\|2020-08 | ···················· | ···················· | ····················· | ···················· | ···················· | ···················· |
| V | 4023 | hCoV-19/Japan/PG-5353/2020\|EPI_ISL_690456\|2020-08 | ···················· | ···················· | ····················· | ···················· | ···················· | ···················· |
| V | 4024 | hCoV-19/Japan/PG-5354/2020\|EPI_ISL_690457\|2020-08 | ···················· | ···················· | ····················· | ···················· | ···················· | ···················· |
| V | 4025 | hCoV-19/Japan/PG-5355/2020\|EPI_ISL_690458\|2020-08 | ···················· | ···················· | ····················· | ···················· | ···················· | ···················· |
| V | 4026 | hCoV-19/Japan/PG-5356/2020\|EPI_ISL_690459\|2020-08 | ···················· | ···················· | ····················· | ···················· | ···················· | ···················· |
| V | 4027 | hCoV-19/Japan/PG-5357/2020\|EPI_ISL_690460\|2020-08 | ···················· | ···················· | ····················· | ···················· | ···················· | ···················· |
| V | 4028 | hCoV-19/Japan/PG-5358/2020\|EPI_ISL_690461\|2020-08 | ···················· | ···················· | ····················· | ···················· | ···················· | ···················· |
| V | 4029 | hCoV-19/Japan/PG-5359/2020\|EPI_ISL_690462\|2020-08 | ···················· | ···················· | ····················· | ···················· | ···················· | ···················· |
| V | 4030 | hCoV-19/Japan/PG-5360/2020\|EPI_ISL_690463\|2020-08 | ···················· | ···················· | ····················· | ···················· | ···················· | ···················· |
| V | 4031 | hCoV-19/Japan/PG-5361/2020\|EPI_ISL_690464\|2020-08 | ···················· | ···················· | ····················· | ···················· | ···················· | ···················· |
| V | 4032 | hCoV-19/Japan/PG-5362/2020\|EPI_ISL_690465\|2020-08 | ···················· | ···················· | ····················· | ···················· | ···················· | ···················· |
| V | 4033 | hCoV-19/Japan/PG-5363/2020\|EPI_ISL_690466\|2020-08 | ···················· | ···················· | ····················· | ···················· | ···················· | ···················· |
| V | 4034 | hCoV-19/Japan/PG-5364/2020\|EPI_ISL_690467\|2020-08 | ···················· | ···················· | ····················· | ···················· | ···················· | ···················· |
| V | 4035 | hCoV-19/Japan/PG-5365/2020\|EPI_ISL_690468\|2020-08 | ···················· | ···················· | ····················· | ···················· | ···················· | ···················· |
| V | 4036 | hCoV-19/Japan/PG-5366/2020\|EPI_ISL_690469\|2020-08 | ···················· | ···················· | ····················· | ···················· | ···················· | ···················· |
| V | 4037 | hCoV-19/Japan/PG-5367/2020\|EPI_ISL_690470\|2020-08 | ···················· | ···················· | ····················· | ···················· | ···················· | ···················· |
| V | 4038 | hCoV-19/Japan/PG-5368/2020\|EPI_ISL_690471\|2020-08 | ···················· | ···················· | ····················· | ···················· | ···················· | ···················· |
| V | 4039 | hCoV-19/Japan/PG-5369/2020\|EPI_ISL_690472\|2020-08 | ···················· | ···················· | ····················· | ···················· | ···················· | ···················· |
| V | 4040 | hCoV-19/Japan/PG-5370/2020\|EPI_ISL_690473\|2020-08 | ···················· | ···················· | ····················· | ···················· | ···················· | ···················· |
| V | 4041 | hCoV-19/Japan/PG-5371/2020\|EPI_ISL_690474\|2020-08 | ···················· | ···················· | ····················· | ···················· | ···················· | ···················· |
| V | 4042 | hCoV-19/Japan/PG-5372/2020\|EPI_ISL_690475\|2020-08 | ···················· | ···················· | ····················· | ···················· | ···················· | ···················· |
| V | 4043 | hCoV-19/Japan/PG-5373/2020\|EPI_ISL_690476\|2020-08 | ···················· | ···················· | ····················· | ···················· | ···················· | ···················· |
| V | 4044 | hCoV-19/Japan/PG-5374/2020\|EPI_ISL_690477\|2020-08 | ···················· | ···················· | ····················· | ···················· | ···················· | ···················· |
| V | 4045 | hCoV-19/Japan/PG-5375/2020\|EPI_ISL_690478\|2020-08 | ···················· | ···················· | ····················· | ···················· | ···················· | ···················· |
| V | 4046 | hCoV-19/Japan/PG-5376/2020\|EPI_ISL_690479\|2020-08 | ···················· | ···················· | ····················· | ···················· | ···················· | ···················· |
| V | 4047 | hCoV-19/Japan/PG-5377/2020\|EPI_ISL_690480\|2020-08 | ···················· | ···················· | ····················· | ···················· | ···················· | ···················· |
| V | 4048 | hCoV-19/Japan/PG-5378/2020\|EPI_ISL_690481\|2020-08 | ···················· | ···················· | ····················· | ···················· | ···················· | ···················· |
| V | 4049 | hCoV-19/Japan/PG-5379/2020\|EPI_ISL_690482\|2020-08 | ···················· | ···················· | ····················· | ···················· | ···················· | ···················· |
| V | 4050 | hCoV-19/Japan/PG-5380/2020\|EPI_ISL_690483\|2020-08 | ···················· | ···················· | ····················· | ···················· | ···················· | ···················· |
| V | 4051 | hCoV-19/Japan/PG-5381/2020\|EPI_ISL_690484\|2020-08 | ···················· | ···················· | ····················· | ···················· | ···················· | ···················· |
| V | 4052 | hCoV-19/Japan/PG-5382/2020\|EPI_ISL_690485\|2020-08 | ···················· | ···················· | ····················· | ···················· | ···················· | ···················· |
| V | 4053 | hCoV-19/Japan/PG-5383/2020\|EPI_ISL_690486\|2020-08 | ···················· | ···················· | ····················· | ···················· | ···················· | ···················· |
| V | 4054 | hCoV-19/Japan/PG-5384/2020\|EPI_ISL_690487\|2020-08 | ···················· | ···················· | ····················· | ···················· | ···················· | ···················· |
| V | 4055 | hCoV-19/Japan/PG-5385/2020\|EPI_ISL_690488\|2020-08 | ···················· | ···················· | ····················· | ···················· | ···················· | ···················· |
| V | 4056 | hCoV-19/Japan/PG-5386/2020\|EPI_ISL_690489\|2020-08 | ···················· | ···················· | ····················· | ···················· | ···················· | ···················· |
| V | 4057 | hCoV-19/Japan/PG-3504/2020\|EPI_ISL_691760\|2020-07 | ···················· | ···················· | ····················· | ···················· | ···················· | ···················· |
| V | 4058 | hCoV-19/Japan/PG-3505/2020\|EPI_ISL_691761\|2020-07 | ···················· | ···················· | ····················· | ···················· | ···················· | ···················· |
| V | 4059 | hCoV-19/Japan/PG-3506/2020\|EPI_ISL_691762\|2020-07 | ···················· | ···················· | ····················· | ···················· | ···················· | ···················· |
| V | 4060 | hCoV-19/Japan/PG-3507/2020\|EPI_ISL_691763\|2020-07 | ···················· | ···················· | ····················· | ···················· | ···················· | ···················· |
| V | 4061 | hCoV-19/Japan/PG-3508/2020\|EPI_ISL_691764\|2020-07 | ···················· | ·A·················· | ····················· | ···················· | ···················· | ···················· |
| V | 4062 | hCoV-19/Japan/PG-3509/2020\|EPI_ISL_691765\|2020-07 | ···················· | ···················· | ····················· | ···················· | ···················· | ···················· |
| V | 4063 | hCoV-19/Japan/PG-3510/2020\|EPI_ISL_691766\|2020-06 | ···················· | ···················· | ····················· | ···················· | ···················· | ···················· |
| V | 4064 | hCoV-19/Japan/PG-3511/2020\|EPI_ISL_691767\|2020-04 | ···················· | ···················· | ····················· | ···················· | ···················· | ···················· |
| V | 4065 | hCoV-19/Japan/PG-3512/2020\|EPI_ISL_691768\|2020-06 | ···················· | ···················· | ····················· | ···················· | ···················· | ···················· |
| V | 4066 | hCoV-19/Japan/PG-3513/2020\|EPI_ISL_691769\|2020-06 | ···················· | ···················· | ····················· | ···················· | ···················· | ···················· |
| V | 4067 | hCoV-19/Japan/PG-3514/2020\|EPI_ISL_691770\|2020-06 | ···················· | ···················· | ····················· | ···················· | ···················· | ···················· |
| V | 4068 | hCoV-19/Japan/PG-3515/2020\|EPI_ISL_691771\|2020-06 | ···················· | ···················· | ····················· | ···················· | ···················· | ···················· |
| V | 4069 | hCoV-19/Japan/PG-3516/2020\|EPI_ISL_691772\|2020-06 | ···················· | ···················· | ··A·················· | ···················· | ···················· | ···················· |
| V | 4070 | hCoV-19/Japan/PG-3517/2020\|EPI_ISL_691773\|2020-06 | ···················· | ···················· | ····················· | ···················· | ···················· | ···················· |

| | | | | | | | | |
|---|---|---|---|---|---|---|---|---|
| V | 4071 | hCoV-19/Japan/PG-3518/2020\|EPI_ISL_691774\|2020-06 | .................... | .................... | ..................... | .................... | .................... | .................... |
| V | 4072 | hCoV-19/Japan/PG-3519/2020\|EPI_ISL_691775\|2020-06 | .................... | .................... | ..................... | .................... | .................... | .................... |
| V | 4073 | hCoV-19/Japan/PG-3520/2020\|EPI_ISL_691776\|2020-06 | .................... | .................... | ..................... | .................... | .................... | .................... |
| V | 4074 | hCoV-19/Japan/PG-3521/2020\|EPI_ISL_691777\|2020-06 | .................... | .................... | ..................... | .................... | .................... | .................... |
| V | 4075 | hCoV-19/Japan/PG-3522/2020\|EPI_ISL_691778\|2020-06 | .................... | .................... | ..................... | .................... | .................... | .................... |
| V | 4076 | hCoV-19/Japan/PG-3523/2020\|EPI_ISL_691779\|2020-06 | .................... | .................... | ..................... | .................... | .................... | .................... |
| V | 4077 | hCoV-19/Japan/PG-3524/2020\|EPI_ISL_691780\|2020-06 | .................... | .................... | ..................... | .................... | .................... | .................... |
| V | 4078 | hCoV-19/Japan/PG-3525/2020\|EPI_ISL_691781\|2020-06 | .................... | .................... | ..................... | .................... | .................... | .................... |
| V | 4079 | hCoV-19/Japan/PG-3526/2020\|EPI_ISL_691782\|2020-06 | .................... | .................... | ..................... | .................... | .................... | .................... |
| V | 4080 | hCoV-19/Japan/PG-3527/2020\|EPI_ISL_691783\|2020-06 | .................... | .................... | ..................... | .................... | .................... | .................... |
| V | 4081 | hCoV-19/Japan/PG-3528/2020\|EPI_ISL_691784\|2020-06 | .................... | .................... | ..................... | .................... | .................... | .................... |
| V | 4082 | hCoV-19/Japan/PG-3529/2020\|EPI_ISL_691785\|2020-06 | .................... | .................... | ..................... | .................... | .................... | .................... |
| V | 4083 | hCoV-19/Japan/PG-3530/2020\|EPI_ISL_691786\|2020-06 | .................... | .................... | ..................... | .................... | .................... | .................... |
| V | 4084 | hCoV-19/Japan/PG-3531/2020\|EPI_ISL_691787\|2020-06 | .................... | .................... | ..................... | .................... | .................... | .................... |
| V | 4085 | hCoV-19/Japan/PG-3532/2020\|EPI_ISL_691788\|2020-06 | .................... | .................... | ..................... | .................... | .................... | .................... |
| V | 4086 | hCoV-19/Japan/PG-3533/2020\|EPI_ISL_691789\|2020-06 | .................... | .................... | ..................... | .................... | .................... | .................... |
| V | 4087 | hCoV-19/Japan/PG-3534/2020\|EPI_ISL_691790\|2020-06 | .................... | .................... | ..................... | .................... | .................... | .................... |
| V | 4088 | hCoV-19/Japan/PG-3535/2020\|EPI_ISL_691791\|2020-06 | .................... | .................... | ..................... | .................... | .................... | .................... |
| V | 4089 | hCoV-19/Japan/PG-3536/2020\|EPI_ISL_691792\|2020-06 | .................... | .................... | ..................... | .................... | .................... | .................... |
| V | 4090 | hCoV-19/Japan/PG-3537/2020\|EPI_ISL_691793\|2020-06 | .................... | .................... | ..................... | .................... | .................... | .................... |
| V | 4091 | hCoV-19/Japan/PG-3538/2020\|EPI_ISL_691794\|2020-06 | .................... | .................... | ..................... | .................... | .................... | .................... |
| V | 4092 | hCoV-19/Japan/PG-3539/2020\|EPI_ISL_691795\|2020-06 | .................... | .................... | ..................... | .................... | .................... | .................... |
| V | 4093 | hCoV-19/Japan/PG-3540/2020\|EPI_ISL_691796\|2020-06 | .................... | .................... | ..................... | .................... | .................... | .................... |
| V | 4094 | hCoV-19/Japan/PG-3541/2020\|EPI_ISL_691797\|2020-06 | .................... | .................... | ..................... | .................... | .................... | .................... |
| V | 4095 | hCoV-19/Japan/PG-3542/2020\|EPI_ISL_691798\|2020-06 | .................... | .................... | ..................... | .................... | .................... | .................... |
| V | 4096 | hCoV-19/Japan/PG-3543/2020\|EPI_ISL_691799\|2020-06 | .................... | .................... | ..................... | .................... | .................... | .................... |
| V | 4097 | hCoV-19/Japan/PG-0930/2020\|EPI_ISL_684540\|2020-04 | .................... | .................... | ..................... | .................... | .................... | .................... |
| V | 4098 | hCoV-19/Japan/PG-0933/2020\|EPI_ISL_684543\|2020-04 | .................... | .................... | ..................... | .................... | .................... | .................... |
| V | 4099 | hCoV-19/Japan/PG-0935/2020\|EPI_ISL_684545\|2020-04 | .................... | .................... | ..................... | .................... | .................... | .................... |
| V | 4100 | hCoV-19/Japan/PG-0936/2020\|EPI_ISL_684546\|2020-04 | .................... | .................... | ..................... | .................... | .................... | .................... |
| V | 4101 | hCoV-19/Japan/PG-0937/2020\|EPI_ISL_684547\|2020-04 | .................... | .................... | ..................... | .................... | .................... | .................... |
| V | 4102 | hCoV-19/Japan/PG-0938/2020\|EPI_ISL_684548\|2020-04 | .................... | .................... | ..................... | .................... | .................... | .................... |
| V | 4103 | hCoV-19/Japan/PG-0939/2020\|EPI_ISL_684549\|2020-04 | .................... | .................... | ..................... | .................... | .................... | .................... |
| V | 4104 | hCoV-19/Japan/PG-0940/2020\|EPI_ISL_684550\|2020-04 | .................... | .................... | ..................... | .................... | .................... | .................... |
| V | 4105 | hCoV-19/Japan/PG-0941/2020\|EPI_ISL_684551\|2020-04 | .................... | .................... | ..................... | .................... | .................... | .................... |
| V | 4106 | hCoV-19/Japan/PG-0942/2020\|EPI_ISL_684552\|2020-04 | .................... | .................... | ..................... | .................... | .................... | .................... |
| V | 4107 | hCoV-19/Japan/PG-0943/2020\|EPI_ISL_684553\|2020-04 | .................... | .................... | ..................... | .................... | .................... | .................... |
| V | 4108 | hCoV-19/Japan/PG-2265/2020\|EPI_ISL_685870\|2020-04 | .................... | .................... | ..................... | .................... | .................... | .................... |
| V | 4109 | hCoV-19/Japan/PG-2267/2020\|EPI_ISL_685872\|2020-04 | .................... | .................... | ..................... | .................... | .................... | .................... |
| V | 4110 | hCoV-19/Japan/PG-2268/2020\|EPI_ISL_685873\|2020-04 | .................... | .................... | ..................... | .................... | .................... | .................... |
| V | 4111 | hCoV-19/Japan/PG-2269/2020\|EPI_ISL_685874\|2020-04 | .................... | .................... | ..................... | .................... | .................... | .................... |
| V | 4112 | hCoV-19/Japan/PG-2270/2020\|EPI_ISL_685875\|2020-04 | .................... | .................... | ..................... | .................... | .................... | .................... |
| V | 4113 | hCoV-19/Japan/PG-2271/2020\|EPI_ISL_685876\|2020-04 | .................... | .................... | ..................... | .................... | .................... | .................... |
| V | 4114 | hCoV-19/Japan/PG-2272/2020\|EPI_ISL_685877\|2020-04 | .................... | .................... | ..................... | .................... | .................... | .................... |
| V | 4115 | hCoV-19/Japan/PG-2273/2020\|EPI_ISL_685878\|2020-04 | .................... | .................... | ..................... | .................... | .................... | .................... |
| V | 4116 | hCoV-19/Japan/PG-2274/2020\|EPI_ISL_685879\|2020-04 | .................... | .................... | ..................... | .................... | .................... | .................... |
| V | 4117 | hCoV-19/Japan/PG-2275/2020\|EPI_ISL_685880\|2020-04 | .................... | .................... | ..................... | .................... | .................... | .................... |
| V | 4118 | hCoV-19/Japan/PG-2276/2020\|EPI_ISL_685881\|2020-04 | .................... | .................... | ..................... | .................... | .................... | .................... |
| V | 4119 | hCoV-19/Japan/PG-2277/2020\|EPI_ISL_685882\|2020-04 | .................... | .................... | ..................... | .................... | .................... | .................... |
| V | 4120 | hCoV-19/Japan/PG-2278/2020\|EPI_ISL_685883\|2020-04 | .................... | .................... | ..................... | .................... | .................... | .................... |
| V | 4121 | hCoV-19/Japan/PG-2279/2020\|EPI_ISL_685884\|2020-04 | .................... | .................... | ..................... | .................... | .................... | .................... |
| V | 4122 | hCoV-19/Japan/PG-2295/2020\|EPI_ISL_685900\|2020-04 | .................... | .................... | ..................... | .................... | .................... | .................... |
| V | 4124 | hCoV-19/Japan/PG-2297/2020\|EPI_ISL_685902\|2020-04 | .................... | .................... | ..................... | .................... | .................... | .................... |
| V | 4125 | hCoV-19/Japan/PG-2298/2020\|EPI_ISL_685903\|2020-04 | .................... | .................... | ..................... | .................... | .................... | .................... |
| V | 4126 | hCoV-19/Japan/PG-2299/2020\|EPI_ISL_685904\|2020-05 | .................... | .................... | ..................... | .................... | .................... | .................... |
| V | 4127 | hCoV-19/Japan/PG-2300/2020\|EPI_ISL_685905\|2020-03 | .................... | .................... | ..................... | .................... | .................... | .................... |
| V | 4128 | hCoV-19/Japan/PG-2301/2020\|EPI_ISL_685906\|2020-04 | .................... | .................... | ..................... | .................... | .................... | .................... |
| V | 4129 | hCoV-19/Japan/PG-2302/2020\|EPI_ISL_685907\|2020-04 | .................... | .................... | ..................... | .................... | .................... | .................... |
| V | 4130 | hCoV-19/Japan/PG-2303/2020\|EPI_ISL_685908\|2020-04 | .................... | .................... | ..................... | .................... | .................... | .................... |
| V | 4131 | hCoV-19/Japan/PG-2304/2020\|EPI_ISL_685909\|2020-05 | .................... | .................... | ..................... | .................... | .................... | .................... |
| V | 4132 | hCoV-19/Japan/PG-2305/2020\|EPI_ISL_685910\|2020-05 | .................... | .................... | ..................... | .................... | .................... | .................... |
| V | 4133 | hCoV-19/Japan/PG-2306/2020\|EPI_ISL_685911\|2020-05 | .................... | .................... | ..................... | .................... | .................... | .................... |
| V | 4134 | hCoV-19/Japan/PG-2307/2020\|EPI_ISL_685912\|2020-05 | .................... | .................... | ..................... | .................... | .................... | .................... |
| V | 4135 | hCoV-19/Japan/PG-2308/2020\|EPI_ISL_685913\|2020-05 | .................... | .................... | ..................... | .................... | .................... | .................... |
| V | 4136 | hCoV-19/Japan/PG-2309/2020\|EPI_ISL_685914\|2020-05 | .................... | .................... | ..................... | .................... | .................... | .................... |
| V | 4137 | hCoV-19/Japan/PG-2310/2020\|EPI_ISL_685915\|2020-05 | .................... | .................... | ..................... | .................... | .................... | .................... |
| V | 4138 | hCoV-19/Japan/PG-2311/2020\|EPI_ISL_685916\|2020-05 | .................... | .................... | ..................... | .................... | .................... | .................... |
| V | 4140 | hCoV-19/Japan/PG-2313/2020\|EPI_ISL_685918\|2020-04 | .................... | .................... | ..................... | .................... | .................... | .................... |
| V | 4141 | hCoV-19/Japan/PG-2314/2020\|EPI_ISL_685919\|2020-04 | .................... | .................... | ..................... | .................... | .................... | .................... |
| V | 4142 | hCoV-19/Japan/PG-7013/2020\|EPI_ISL_688118\|2020-08 | .................... | .................... | ..................... | .................... | .................... | .................... |
| V | 4143 | hCoV-19/Japan/PG-7014/2020\|EPI_ISL_688119\|2020-08 | .................... | .................... | ..................... | .................... | .................... | .................... |
| V | 4144 | hCoV-19/Japan/PG-7015/2020\|EPI_ISL_688120\|2020-08 | .................... | .................... | ..................... | .................... | .................... | .................... |
| V | 4145 | hCoV-19/Japan/PG-7016/2020\|EPI_ISL_688121\|2020-08 | .................... | .................... | ..................... | .................... | .................... | .................... |
| V | 4146 | hCoV-19/Japan/PG-7017/2020\|EPI_ISL_688122\|2020-08 | .................... | .................... | ..................... | .................... | .................... | .................... |
| V | 4147 | hCoV-19/Japan/PG-7018/2020\|EPI_ISL_688123\|2020-08 | .................... | .................... | ..................... | .................... | .................... | .................... |
| V | 4148 | hCoV-19/Japan/PG-7019/2020\|EPI_ISL_688124\|2020-08 | .................... | .................... | ..................... | .................... | .................... | .................... |
| V | 4149 | hCoV-19/Japan/PG-7020/2020\|EPI_ISL_688125\|2020-08 | .................... | .................... | ..................... | .................... | .................... | .................... |
| V | 4150 | hCoV-19/Japan/PG-7021/2020\|EPI_ISL_688126\|2020-08 | .................... | .................... | ..................... | .................... | .................... | .................... |
| V | 4151 | hCoV-19/Japan/PG-7022/2020\|EPI_ISL_688127\|2020-08 | .................... | .................... | ..................... | .................... | .................... | .................... |
| V | 4152 | hCoV-19/Japan/PG-7023/2020\|EPI_ISL_688128\|2020-08 | .................... | .................... | ..................... | .................... | .................... | .................... |

V 4153 hCoV-19/Japan/PG-7024/2020|EPI_ISL_688129|2020-08 .................... .................... ..................... .................... .................... ....................
V 4155 hCoV-19/Japan/PG-7026/2020|EPI_ISL_688131|2020-08 .................... .................... ..................... .................... .................... ....................
V 4156 hCoV-19/Japan/PG-7027/2020|EPI_ISL_688132|2020-08 .................... .................... ..................... .................... .................... ....................
V 4157 hCoV-19/Japan/PG-7028/2020|EPI_ISL_688133|2020-08 .................... .................... ..................... .................... .................... ....................
V 4158 hCoV-19/Japan/PG-7029/2020|EPI_ISL_688134|2020-08 .................... .................... ..................... .................... .................... ....................
V 4159 hCoV-19/Japan/PG-7030/2020|EPI_ISL_688135|2020-08 .................... .................... ..................... .................... .................... ....................
V 4160 hCoV-19/Japan/PG-7031/2020|EPI_ISL_688136|2020-08 .................... .................... ..................... .................... .................... ....................
V 4161 hCoV-19/Japan/PG-7032/2020|EPI_ISL_688137|2020-08 .................... .................... ..................... .................... .................... ....................
V 4162 hCoV-19/Japan/PG-7033/2020|EPI_ISL_688138|2020-08 .................... .................... ..................... .................... .................... ....................
V 4163 hCoV-19/Japan/PG-7034/2020|EPI_ISL_688139|2020-08 .................... .................... ..................... .................... .................... ....................
V 4164 hCoV-19/Japan/PG-7035/2020|EPI_ISL_688140|2020-08 .................... .................... ..................... .................... .................... ....................
V 4165 hCoV-19/Japan/PG-7036/2020|EPI_ISL_688141|2020-08 .................... .................... ..................... .................... .................... ....................
V 4166 hCoV-19/Japan/PG-7037/2020|EPI_ISL_688142|2020-08 .................... .................... ..................... .................... .................... ....................
V 4167 hCoV-19/Japan/PG-7038/2020|EPI_ISL_688143|2020-08 .................... .................... ..................... .................... .................... ....................
V 4168 hCoV-19/Japan/PG-7039/2020|EPI_ISL_688144|2020-08 .................... .................... ..................... .................... .................... ....................
V 4169 hCoV-19/Japan/PG-7040/2020|EPI_ISL_688145|2020-08 .................... .................... ..................... .................... .................... ....................
V 4170 hCoV-19/Japan/PG-7041/2020|EPI_ISL_688146|2020-08 .................... .................... ..................... .................... .................... ....................
V 4171 hCoV-19/Japan/PG-7042/2020|EPI_ISL_688147|2020-08 .................... .................... ..................... .................... .................... ....................
V 4172 hCoV-19/Japan/PG-7043/2020|EPI_ISL_688148|2020-08 .................... .................... ..................... .................... .................... ....................
V 4174 hCoV-19/Japan/PG-7045/2020|EPI_ISL_688150|2020-08 .................... .................... ..................... .................... .................... ....................
V 4175 hCoV-19/Japan/PG-7046/2020|EPI_ISL_688151|2020-08 .................... .................... ..................... .................... .................... ....................
V 4176 hCoV-19/Japan/PG-7047/2020|EPI_ISL_688152|2020-08 .................... .................... ..................... .................... .................... ....................
V 4177 hCoV-19/Japan/PG-7048/2020|EPI_ISL_688153|2020-08 .................... .................... ..................... .................... .................... ....................
V 4178 hCoV-19/Japan/PG-7049/2020|EPI_ISL_688154|2020-08 .................... .................... ..................... .................... .................... ....................
V 4179 hCoV-19/Japan/PG-7050/2020|EPI_ISL_688155|2020-08 .................... .................... ..................... .................... .................... ....................
V 4180 hCoV-19/Japan/PG-7051/2020|EPI_ISL_688156|2020-08 .................... .................... ..................... .................... .................... ....................
V 4181 hCoV-19/Japan/PG-7052/2020|EPI_ISL_688157|2020-08 .................... .................... ..................... .................... .................... ....................
V 4182 hCoV-19/Japan/PG-7053/2020|EPI_ISL_688158|2020-08 .................... .................... ..................... .................... .................... ....................
V 4183 hCoV-19/Japan/PG-7054/2020|EPI_ISL_688159|2020-08 .................... .................... ..................... .................... .................... ....................
V 4184 hCoV-19/Japan/PG-7055/2020|EPI_ISL_688160|2020-08 .................... .................... ..................... .................... .................... ....................
V 4185 hCoV-19/Japan/PG-7056/2020|EPI_ISL_688161|2020-08 .................... .................... ..................... .................... .................... ....................
V 4186 hCoV-19/Japan/PG-7057/2020|EPI_ISL_688162|2020-08 .................... .................... ..................... .................... .................... ....................
V 4187 hCoV-19/Japan/PG-7058/2020|EPI_ISL_688163|2020-08 .................... .................... ..................... .................... .................... ....................
V 4188 hCoV-19/Japan/PG-7059/2020|EPI_ISL_688164|2020-08 .................... .................... ..................... .................... .................... ....................
V 4189 hCoV-19/Japan/PG-7060/2020|EPI_ISL_688165|2020-08 .................... .................... ..................... .................... .................... ....................
V 4190 hCoV-19/Japan/PG-7061/2020|EPI_ISL_688166|2020-08 .................... .................... ..................... .................... .................... ....................
V 4191 hCoV-19/Japan/PG-7062/2020|EPI_ISL_688167|2020-08 .................... .................... ..................... .................... .................... ....................
V 4192 hCoV-19/Japan/PG-7063/2020|EPI_ISL_688168|2020-08 .................... .................... ..................... .................... .................... ....................
V 4193 hCoV-19/Japan/PG-7064/2020|EPI_ISL_688169|2020-08 .................... .................... ..................... .................... .................... ....................
V 4194 hCoV-19/Japan/PG-7065/2020|EPI_ISL_688170|2020-08 .................... .................... ..................... .................... .................... ....................
V 4195 hCoV-19/Japan/PG-7066/2020|EPI_ISL_688171|2020-08 .................... .................... ..................... .................... .................... ....................
V 4196 hCoV-19/Japan/PG-7067/2020|EPI_ISL_688172|2020-08 .................... .................... ..................... .................... .................... ....................
V 4197 hCoV-19/Japan/PG-7068/2020|EPI_ISL_688173|2020-08 .................... .................... ..................... .................... .................... ....................
V 4198 hCoV-19/Japan/PG-7069/2020|EPI_ISL_688174|2020-08 .................... .................... ..................... .................... .................... ....................
V 4199 hCoV-19/Japan/PG-7070/2020|EPI_ISL_688175|2020-08 .................... .................... ..................... .................... .................... ....................
V 4200 hCoV-19/Japan/PG-7071/2020|EPI_ISL_688176|2020-08 .................... .................... ..................... .................... .................... ....................
V 4201 hCoV-19/Japan/PG-7072/2020|EPI_ISL_688177|2020-08 .................... .................... ..................... .................... .................... ....................
V 4202 hCoV-19/Japan/PG-7073/2020|EPI_ISL_688178|2020-08 .................... .................... ..................... .................... .................... ....................
V 4203 hCoV-19/Japan/PG-7074/2020|EPI_ISL_688179|2020-08 .................... .................... ..................... .................... .................... ....................
V 4204 hCoV-19/Japan/PG-7075/2020|EPI_ISL_688180|2020-08 .................... .................... ..................... .................... .................... ....................
V 4205 hCoV-19/Japan/PG-7076/2020|EPI_ISL_688181|2020-08 .................... .................... ..................... .................... .................... ....................
V 4206 hCoV-19/Japan/PG-7077/2020|EPI_ISL_688182|2020-08 .................... .................... ..................... .................... .................... ....................
V 4207 hCoV-19/Japan/PG-7078/2020|EPI_ISL_688183|2020-08 .................... .................... ..................... .................... .................... ....................
V 4208 hCoV-19/Japan/PG-7079/2020|EPI_ISL_688184|2020-08 .................... .................... ..................... .................... .................... ....................
V 4209 hCoV-19/Japan/PG-7080/2020|EPI_ISL_688185|2020-08 .................... .................... ..................... .................... .................... ....................
V 4211 hCoV-19/Japan/PG-7082/2020|EPI_ISL_688187|2020-08 .................... .................... ..................... .................... .................... ....................
V 4212 hCoV-19/Japan/PG-7083/2020|EPI_ISL_688188|2020-08 .................... .................... ..................... .................... .................... ....................
V 4213 hCoV-19/Japan/PG-7084/2020|EPI_ISL_688189|2020-08 .................... .................... ..................... .................... .................... ....................
V 4214 hCoV-19/Japan/PG-7085/2020|EPI_ISL_688190|2020-08 .................... .................... ..................... .................... .................... ....................
V 4215 hCoV-19/Japan/PG-7086/2020|EPI_ISL_688191|2020-08 .................... .................... ..................... .................... .................... ....................
V 4216 hCoV-19/Japan/PG-7087/2020|EPI_ISL_688192|2020-08 .................... .................... ..................... .................... .................... ....................
V 4217 hCoV-19/Japan/PG-7088/2020|EPI_ISL_688193|2020-08 .................... .................... ..................... .................... .................... ....................
V 4218 hCoV-19/Japan/PG-7089/2020|EPI_ISL_688194|2020-08 .................... .................... ..................... .................... .................... ....................
V 4219 hCoV-19/Japan/PG-7090/2020|EPI_ISL_688195|2020-08 .................... .................... ..................... .................... .................... ....................
V 4220 hCoV-19/Japan/PG-7091/2020|EPI_ISL_688196|2020-08 .................... .................... ..................... .................... .................... ....................
V 4221 hCoV-19/Japan/PG-7092/2020|EPI_ISL_688197|2020-08 .................... .................... ..................... .................... .................... ....................
V 4222 hCoV-19/Japan/PG-7093/2020|EPI_ISL_688198|2020-08 .................... .................... ..................... .................... .................... ....................
V 4223 hCoV-19/Japan/PG-7094/2020|EPI_ISL_688199|2020-08 .................... .................... ..................... .................... .................... ....................
V 4224 hCoV-19/Japan/PG-6338/2020|EPI_ISL_689449|2020-08 .................... .................... ..................... .................... .................... ....................
V 4225 hCoV-19/Japan/PG-6339/2020|EPI_ISL_689450|2020-08 .................... .................... ..................... .................... .................... ....................
V 4226 hCoV-19/Japan/PG-6340/2020|EPI_ISL_689451|2020-08 .................... .................... ..................... .................... .................... ....................
V 4227 hCoV-19/Japan/PG-6341/2020|EPI_ISL_689452|2020-08 .................... .................... ..................... .................... .................... ....................
V 4228 hCoV-19/Japan/PG-6342/2020|EPI_ISL_689453|2020-08 .................... .................... ..................... .................... .................... ....................
V 4229 hCoV-19/Japan/PG-6343/2020|EPI_ISL_689454|2020-08 .................... .................... ..................... .................... .................... ....................
V 4230 hCoV-19/Japan/PG-6344/2020|EPI_ISL_689455|2020-08 .................... .................... ..................... .................... .................... ....................
V 4231 hCoV-19/Japan/PG-6345/2020|EPI_ISL_689456|2020-08 .................... .................... ..................... .................... .................... ....................
V 4232 hCoV-19/Japan/PG-6346/2020|EPI_ISL_689457|2020-08 .................... .................... ..................... .................... .................... ....................
V 4233 hCoV-19/Japan/PG-6347/2020|EPI_ISL_689458|2020-08 .................... .................... ..................... .................... .................... ....................
V 4234 hCoV-19/Japan/PG-6348/2020|EPI_ISL_689459|2020-08 .................... .................... ..................... .................... .................... ....................
V 4235 hCoV-19/Japan/PG-6349/2020|EPI_ISL_689460|2020-08 .................... .................... ..................... .................... .................... ....................

| | | | | | | | | |
|---|---|---|---|---|---|---|---|---|
| V | 4236 | hCoV-19/Japan/PG-6350/2020\|EPI_ISL_689461\|2020-08 | .................... | .................... | .................... | .................... | .................... | .................... |
| V | 4237 | hCoV-19/Japan/PG-6351/2020\|EPI_ISL_689462\|2020-08 | .................... | .................... | .................... | .................... | .................... | .................... |
| V | 4238 | hCoV-19/Japan/PG-6352/2020\|EPI_ISL_689463\|2020-08 | .................... | .................... | .................... | .................... | .................... | .................... |
| V | 4239 | hCoV-19/Japan/PG-6353/2020\|EPI_ISL_689464\|2020-08 | .................... | .................... | .................... | .................... | .................... | .................... |
| V | 4240 | hCoV-19/Japan/PG-6354/2020\|EPI_ISL_689465\|2020-08 | .................... | .................... | .................... | .................... | .................... | .................... |
| V | 4241 | hCoV-19/Japan/PG-6355/2020\|EPI_ISL_689466\|2020-08 | .................... | .................... | .................... | .................... | .................... | .................... |
| V | 4242 | hCoV-19/Japan/PG-6356/2020\|EPI_ISL_689467\|2020-08 | .................... | .................... | .................... | .................... | .................... | .................... |
| V | 4243 | hCoV-19/Japan/PG-6357/2020\|EPI_ISL_689468\|2020-08 | .................... | .................... | .................... | .................... | .................... | .................... |
| V | 4244 | hCoV-19/Japan/PG-6358/2020\|EPI_ISL_689469\|2020-08 | .................... | .................... | .................... | .................... | .................... | .................... |
| V | 4245 | hCoV-19/Japan/PG-6359/2020\|EPI_ISL_689470\|2020-08 | .................... | .................... | .................... | .................... | .................... | .................... |
| V | 4246 | hCoV-19/Japan/PG-6360/2020\|EPI_ISL_689471\|2020-08 | .................... | .................... | .................... | .................... | .................... | .................... |
| V | 4247 | hCoV-19/Japan/PG-6361/2020\|EPI_ISL_689472\|2020-08 | .................... | .................... | .................... | .................... | .................... | .................... |
| V | 4248 | hCoV-19/Japan/PG-6362/2020\|EPI_ISL_689473\|2020-08 | .................... | .................... | .................... | .................... | .................... | .................... |
| V | 4249 | hCoV-19/Japan/PG-6363/2020\|EPI_ISL_689474\|2020-08 | .................... | .................... | .................... | .................... | .................... | .................... |
| V | 4250 | hCoV-19/Japan/PG-6364/2020\|EPI_ISL_689475\|2020-08 | .................... | .................... | .................... | .................... | .................... | .................... |
| V | 4251 | hCoV-19/Japan/PG-6365/2020\|EPI_ISL_689476\|2020-08 | .................... | .................... | .................... | .................... | .................... | .................... |
| V | 4252 | hCoV-19/Japan/PG-6366/2020\|EPI_ISL_689477\|2020-08 | .................... | .................... | .................... | .................... | .................... | .................... |
| V | 4253 | hCoV-19/Japan/PG-6367/2020\|EPI_ISL_689478\|2020-08 | .................... | .................... | .................... | .................... | .................... | .................... |
| V | 4254 | hCoV-19/Japan/PG-6368/2020\|EPI_ISL_689479\|2020-08 | .................... | .................... | .................... | .................... | .................... | .................... |
| V | 4255 | hCoV-19/Japan/PG-6369/2020\|EPI_ISL_689480\|2020-08 | .................... | .................... | .................... | .................... | .................... | .................... |
| V | 4256 | hCoV-19/Japan/PG-6370/2020\|EPI_ISL_689481\|2020-08 | .................... | .................... | .................... | .................... | .................... | .................... |
| V | 4257 | hCoV-19/Japan/PG-6371/2020\|EPI_ISL_689482\|2020-08 | .................... | .................... | .................... | .................... | .................... | .................... |
| V | 4258 | hCoV-19/Japan/PG-6372/2020\|EPI_ISL_689483\|2020-08 | .................... | .................... | .................... | .................... | .................... | .................... |
| V | 4259 | hCoV-19/Japan/PG-6373/2020\|EPI_ISL_689484\|2020-08 | .................... | .................... | .................... | .................... | .................... | .................... |
| V | 4260 | hCoV-19/Japan/PG-6374/2020\|EPI_ISL_689485\|2020-08 | .................... | .................... | .................... | .................... | .................... | .................... |
| V | 4261 | hCoV-19/Japan/PG-6375/2020\|EPI_ISL_689486\|2020-08 | .................... | .................... | .................... | .................... | .................... | .................... |
| V | 4262 | hCoV-19/Japan/PG-6376/2020\|EPI_ISL_689487\|2020-08 | .................... | .................... | .................... | .................... | .................... | .................... |
| V | 4263 | hCoV-19/Japan/PG-6377/2020\|EPI_ISL_689488\|2020-08 | .................... | .................... | .................... | .................... | .................... | .................... |
| V | 4264 | hCoV-19/Japan/PG-6379/2020\|EPI_ISL_689489\|2020-08 | .................... | .................... | .................... | .................... | .................... | .................... |
| V | 4265 | hCoV-19/Japan/PG-6380/2020\|EPI_ISL_689490\|2020-08 | .................... | .................... | .................... | .................... | .................... | .................... |
| V | 4266 | hCoV-19/Japan/PG-6381/2020\|EPI_ISL_689491\|2020-08 | .................... | .................... | .................... | .................... | .................... | .................... |
| V | 4267 | hCoV-19/Japan/PG-6382/2020\|EPI_ISL_689492\|2020-08 | .................... | .................... | .................... | .................... | .................... | .................... |
| V | 4268 | hCoV-19/Japan/PG-6384/2020\|EPI_ISL_689493\|2020-08 | .................... | .................... | .................... | .................... | .................... | .................... |
| V | 4269 | hCoV-19/Japan/PG-6385/2020\|EPI_ISL_689494\|2020-08 | .................... | .................... | .................... | .................... | .................... | .................... |
| V | 4270 | hCoV-19/Japan/PG-6386/2020\|EPI_ISL_689495\|2020-08 | .................... | .................... | .................... | .................... | .................... | .................... |
| V | 4271 | hCoV-19/Japan/PG-6387/2020\|EPI_ISL_689496\|2020-08 | .................... | .................... | .................... | .................... | .................... | .................... |
| V | 4272 | hCoV-19/Japan/PG-6391/2020\|EPI_ISL_689497\|2020-08 | .................... | .................... | .................... | .................... | .................... | .................... |
| V | 4273 | hCoV-19/Japan/PG-6392/2020\|EPI_ISL_689498\|2020-08 | .................... | .................... | .................... | .................... | .................... | .................... |
| V | 4274 | hCoV-19/Japan/PG-6393/2020\|EPI_ISL_689499\|2020-08 | .................... | .................... | .................... | .................... | .................... | .................... |
| V | 4275 | hCoV-19/Japan/PG-5297/2020\|EPI_ISL_690400\|2020-08 | .................... | .................... | .................... | .................... | .................... | .................... |
| V | 4276 | hCoV-19/Japan/PG-5298/2020\|EPI_ISL_690401\|2020-08 | .................... | .................... | .................... | .................... | .................... | .................... |
| V | 4277 | hCoV-19/Japan/PG-5299/2020\|EPI_ISL_690402\|2020-08 | .................... | .................... | .................... | .................... | .................... | .................... |
| V | 4278 | hCoV-19/Japan/PG-5300/2020\|EPI_ISL_690403\|2020-08 | .................... | .................... | .................... | .................... | .................... | .................... |
| V | 4279 | hCoV-19/Japan/PG-5301/2020\|EPI_ISL_690404\|2020-08 | .................... | .................... | .................... | .................... | .................... | .................... |
| V | 4280 | hCoV-19/Japan/PG-5302/2020\|EPI_ISL_690405\|2020-08 | .................... | .................... | .................... | .................... | .................... | .................... |
| V | 4281 | hCoV-19/Japan/PG-5303/2020\|EPI_ISL_690406\|2020-08 | .................... | .................... | .................... | .................... | .................... | .................... |
| V | 4282 | hCoV-19/Japan/PG-5304/2020\|EPI_ISL_690407\|2020-08 | .................... | .................... | .................... | .................... | .................... | .................... |
| V | 4283 | hCoV-19/Japan/PG-5305/2020\|EPI_ISL_690408\|2020-08 | .................... | .................... | .................... | .................... | .................... | .................... |
| V | 4284 | hCoV-19/Japan/PG-5306/2020\|EPI_ISL_690409\|2020-08 | .................... | .................... | .................... | .................... | .................... | .................... |
| V | 4285 | hCoV-19/Japan/PG-5307/2020\|EPI_ISL_690410\|2020-08 | .................... | .................... | .................... | .................... | .................... | .................... |
| V | 4286 | hCoV-19/Japan/PG-5308/2020\|EPI_ISL_690411\|2020-08 | .................... | .................... | .................... | .................... | .................... | .................... |
| V | 4287 | hCoV-19/Japan/PG-5309/2020\|EPI_ISL_690412\|2020-08 | .................... | .................... | .................... | .................... | .................... | .................... |
| V | 4288 | hCoV-19/Japan/PG-5310/2020\|EPI_ISL_690413\|2020-08 | .................... | .................... | .................... | .................... | .................... | .................... |
| V | 4289 | hCoV-19/Japan/PG-5311/2020\|EPI_ISL_690414\|2020-08 | .................... | .................... | .................... | .................... | .................... | .................... |
| V | 4290 | hCoV-19/Japan/PG-5312/2020\|EPI_ISL_690415\|2020-08 | .................... | .................... | .................... | .................... | .................... | .................... |
| V | 4291 | hCoV-19/Japan/PG-5313/2020\|EPI_ISL_690416\|2020-08 | .................... | .................... | .................... | .................... | .................... | .................... |
| V | 4292 | hCoV-19/Japan/PG-5314/2020\|EPI_ISL_690417\|2020-08 | .................... | .................... | .................... | .................... | .................... | .................... |
| V | 4293 | hCoV-19/Japan/PG-5315/2020\|EPI_ISL_690418\|2020-08 | .................... | .................... | .................... | .................... | .................... | .................... |
| V | 4294 | hCoV-19/Japan/PG-5316/2020\|EPI_ISL_690419\|2020-08 | .................... | .................... | .................... | .................... | .................... | .................... |
| V | 4296 | hCoV-19/Japan/PG-5318/2020\|EPI_ISL_690421\|2020-08 | .................... | .................... | .................... | .................... | .................... | .................... |
| V | 4298 | hCoV-19/Japan/PG-5320/2020\|EPI_ISL_690423\|2020-08 | .................... | .................... | .................... | .................... | .................... | .................... |
| V | 4299 | hCoV-19/Japan/PG-5321/2020\|EPI_ISL_690424\|2020-08 | .................... | .................... | .................... | .................... | .................... | .................... |
| V | 4300 | hCoV-19/Japan/PG-5322/2020\|EPI_ISL_690425\|2020-08 | .................... | .................... | .................... | .................... | .................... | .................... |
| V | 4301 | hCoV-19/Japan/PG-5323/2020\|EPI_ISL_690426\|2020-08 | .................... | .................... | .................... | .................... | .................... | .................... |
| V | 4302 | hCoV-19/Japan/PG-5324/2020\|EPI_ISL_690427\|2020-08 | .................... | .................... | .................... | .................... | .................... | .................... |
| V | 4303 | hCoV-19/Japan/PG-5325/2020\|EPI_ISL_690428\|2020-08 | .................... | .................... | .................... | .................... | .................... | .................... |
| V | 4304 | hCoV-19/Japan/PG-5326/2020\|EPI_ISL_690429\|2020-08 | .................... | .................... | .................... | .................... | .................... | .................... |
| V | 4305 | hCoV-19/Japan/PG-5387/2020\|EPI_ISL_690490\|2020-08 | .................... | .................... | .................... | .................... | .................... | .................... |
| V | 4306 | hCoV-19/Japan/PG-5388/2020\|EPI_ISL_690491\|2020-08 | .................... | .................... | .................... | .................... | .................... | .................... |
| V | 4307 | hCoV-19/Japan/PG-5389/2020\|EPI_ISL_690492\|2020-08 | .................... | .................... | .................... | .................... | .................... | .................... |
| V | 4308 | hCoV-19/Japan/PG-5390/2020\|EPI_ISL_690493\|2020-08 | .................... | .................... | .................... | .................... | .................... | .................... |
| V | 4309 | hCoV-19/Japan/PG-5391/2020\|EPI_ISL_690494\|2020-08 | .................... | .................... | .................... | .................... | .................... | .................... |
| V | 4310 | hCoV-19/Japan/PG-5392/2020\|EPI_ISL_690495\|2020-08 | .................... | .................... | .................... | .................... | .................... | .................... |
| V | 4311 | hCoV-19/Japan/PG-5393/2020\|EPI_ISL_690496\|2020-08 | .................... | .................... | .................... | .................... | .................... | .................... |
| V | 4312 | hCoV-19/Japan/PG-5394/2020\|EPI_ISL_690497\|2020-08 | .................... | .................... | .................... | .................... | .................... | .................... |
| V | 4313 | hCoV-19/Japan/PG-5395/2020\|EPI_ISL_690498\|2020-08 | .................... | .................... | .................... | .................... | .................... | .................... |
| V | 4314 | hCoV-19/Japan/PG-5396/2020\|EPI_ISL_690499\|2020-08 | .................... | .................... | .................... | .................... | .................... | .................... |
| V | 4315 | hCoV-19/Spain/MD-ISCIII-2014585/2020\|EPI_ISL_69170 | .................... | .................... | .................... | .................... | .................... | .................... |
| V | 4316 | hCoV-19/Spain/MD-ISCIII-2014592/2020\|EPI_ISL_69170 | .................... | .................... | .................... | .................... | .................... | .................... |
| V | 4317 | hCoV-19/Spain/MD-ISCIII-2014599/2020\|EPI_ISL_69170 | .................... | .................... | .................... | .................... | .................... | .................... |

| | | | | | | | |
|---|---|---|---|---|---|---|---|
| V | 4318 hCoV-19/Spain/MD-ISCIII-2014601/2020\|EPI_ISL_69170 | .................... | .................... | .................... | .................... | .................... | .................... |
| V | 4319 hCoV-19/Spain/MD-ISCIII-2014603/2020\|EPI_ISL_69170 | .................... | .................... | .................... | .................... | .................... | .................... |
| V | 4320 hCoV-19/Spain/MD-ISCIII-2014615/2020\|EPI_ISL_69170 | .................... | .................... | .................... | .................... | .................... | .................... |
| V | 4321 hCoV-19/Spain/MD-ISCIII-2014616/2020\|EPI_ISL_69170 | .................... | .................... | .................... | .................... | .................... | .................... |
| V | 4322 hCoV-19/Spain/MD-ISCIII-2014617/2020\|EPI_ISL_69170 | .................... | .................... | .................... | .................... | .................... | .................... |
| V | 4323 hCoV-19/Spain/MD-ISCIII-2014623/2020\|EPI_ISL_69170 | .................... | .................... | .................... | .................... | .................... | .................... |
| V | 4324 hCoV-19/Spain/MD-ISCIII-2014624/2020\|EPI_ISL_69170 | .................... | .................... | .................... | .................... | .................... | .................... |
| V | 4325 hCoV-19/Spain/MD-ISCIII-2014625/2020\|EPI_ISL_69171 | .................... | .................... | .................... | .................... | .................... | .................... |
| V | 4326 hCoV-19/Spain/MD-ISCIII-2014627/2020\|EPI_ISL_69171 | .................... | .................... | .................... | .................... | .................... | .................... |
| V | 4327 hCoV-19/Spain/MD-ISCIII-2014628/2020\|EPI_ISL_69171 | .................... | .................... | .................... | .................... | .................... | .................... |
| V | 4328 hCoV-19/Spain/MD-ISCIII-2014630/2020\|EPI_ISL_69171 | .................... | .................... | .................... | .................... | .................... | .................... |
| V | 4329 hCoV-19/Spain/MD-ISCIII-2014631/2020\|EPI_ISL_69171 | .................... | .................... | .................... | .................... | .................... | .................... |
| V | 4330 hCoV-19/Spain/MD-ISCIII-2014632/2020\|EPI_ISL_69171 | .................... | .................... | .................... | .................... | .................... | .................... |
| V | 4331 hCoV-19/Spain/MD-ISCIII-2014656/2020\|EPI_ISL_69171 | .................... | .................... | .................... | .................... | .................... | .................... |
| V | 4332 hCoV-19/Spain/MD-ISCIII-2014662/2020\|EPI_ISL_69171 | .................... | .................... | .................... | .................... | .................... | .................... |
| V | 4333 hCoV-19/Spain/MD_ISCIII-2014663/2020\|EPI_ISL_69171 | .................... | .................... | .................... | .................... | .................... | .................... |
| V | 4334 hCoV-19/Spain/MD-ISCIII-2014664/2020\|EPI_ISL_69171 | .................... | .................... | .................... | .................... | .................... | .................... |
| V | 4335 hCoV-19/Spain/MD-ISCIII-2014669/2020\|EPI_ISL_69172 | .................... | .................... | .................... | .................... | .................... | .................... |
| V | 4336 hCoV-19/Spain/MD-ISCIII-2014670/2020\|EPI_ISL_69172 | .................... | .................... | .................... | .................... | .................... | .................... |
| V | 4337 hCoV-19/Spain/MD-ISCIII-2014780/2020\|EPI_ISL_69172 | .................... | .................... | .................... | .................... | .................... | .................... |
| V | 4338 hCoV-19/Spain/MD-ISCIII-2014822/2020\|EPI_ISL_69172 | .................... | .................... | .................... | .................... | .................... | .................... |
| V | 4339 hCoV-19/Spain/MD-ISCIII-2014824/2020\|EPI_ISL_69172 | .................... | .................... | .................... | .................... | .................... | .................... |
| V | 4340 hCoV-19/Spain/MD-ISCIII-2014825/2020\|EPI_ISL_69172 | .................... | .................... | .................... | .................... | .................... | .................... |
| V | 4341 hCoV-19/Spain/MD-ISCIII-2014828/2020\|EPI_ISL_69172 | .................... | .................... | .................... | .................... | .................... | .................... |
| V | 4342 hCoV-19/Spain/MD-ISCIII-2014829/2020\|EPI_ISL_69172 | .................... | .................... | .................... | .................... | .................... | .................... |
| V | 4343 hCoV-19/Spain/MD-ISCIII-2014937/2020\|EPI_ISL_69172 | .................... | .................... | .................... | .................... | .................... | .................... |
| V | 4344 hCoV-19/Spain/MD-ISCIII-2014833/2020\|EPI_ISL_69172 | .................... | .................... | .................... | .................... | .................... | .................... |
| V | 4345 hCoV-19/Spain/MD-ISCIII-2015114/2020\|EPI_ISL_69173 | .................... | .................... | .................... | .................... | .................... | .................... |
| V | 4346 hCoV-19/Spain/MD-ISCIII-2016006/2020\|EPI_ISL_69173 | .................... | .................... | .................... | .................... | .................... | .................... |
| V | 4347 hCoV-19/Spain/MD-ISCIII-2016128/2020\|EPI_ISL_69173 | .................... | .................... | .................... | .................... | .................... | .................... |
| V | 4348 hCoV-19/Japan/PG-4364/2020\|EPI_ISL_691744\|2020-07 | .................... | .................... | .................... | .................... | .................... | .................... |
| V | 4349 hCoV-19/Japan/PG-4106/2020\|EPI_ISL_691748\|2020-06 | .................... | .................... | .................... | .................... | .................... | .................... |
| V | 4350 hCoV-19/Japan/PG-3610/2020\|EPI_ISL_691752\|2020-07 | .................... | .................... | .................... | .................... | .................... | .................... |
| V | 4351 hCoV-19/Japan/PG-4432/2020\|EPI_ISL_691753\|2020-07 | .................... | .................... | .................... | .................... | .................... | .................... |
| V | 4352 hCoV-19/Japan/PG-3503/2020\|EPI_ISL_691759\|2020-07 | .................... | .................... | .................... | .................... | .................... | .................... |
| V | 4353 hCoV-19/Japan/PG-0890/2020\|EPI_ISL_684500\|2020-03 | .................... | .................... | .................... | .................... | .................... | .................... |
| V | 4354 hCoV-19/Japan/PG-0891/2020\|EPI_ISL_684501\|2020-03 | .................... | .................... | .................... | .................... | .................... | .................... |
| V | 4355 hCoV-19/Japan/PG-0892/2020\|EPI_ISL_684502\|2020-04 | .................... | .................... | .................... | .................... | .................... | .................... |
| V | 4356 hCoV-19/Japan/PG-0893/2020\|EPI_ISL_684503\|2020-04 | .................... | .................... | .................... | .................... | .................... | .................... |
| V | 4357 hCoV-19/Japan/PG-0894/2020\|EPI_ISL_684504\|2020-04 | .................... | .................... | .................... | .................... | .................... | .................... |
| V | 4358 hCoV-19/Japan/PG-0895/2020\|EPI_ISL_684505\|2020-04 | .................... | .................... | .................... | .................... | .................... | .................... |
| V | 4359 hCoV-19/Japan/PG-0896/2020\|EPI_ISL_684506\|2020-03 | .................... | .................... | .................... | .................... | .................... | .................... |
| V | 4360 hCoV-19/Japan/PG-0897/2020\|EPI_ISL_684507\|2020-03 | .................... | .................... | .................... | .................... | .................... | .................... |
| V | 4361 hCoV-19/Japan/PG-0898/2020\|EPI_ISL_684508\|2020-03 | .................... | .................... | .................... | .................... | .................... | .................... |
| V | 4362 hCoV-19/Japan/PG-0899/2020\|EPI_ISL_684509\|2020-03 | .................... | .................... | .................... | .................... | .................... | .................... |
| V | 4363 hCoV-19/Japan/PG-0900/2020\|EPI_ISL_684510\|2020-03 | .................... | .................... | .................... | .................... | .................... | .................... |
| V | 4364 hCoV-19/Japan/PG-0901/2020\|EPI_ISL_684511\|2020-03 | .................... | .................... | .................... | .................... | .................... | .................... |
| V | 4365 hCoV-19/Japan/PG-0902/2020\|EPI_ISL_684512\|2020-03 | .................... | .................... | .................... | .................... | .................... | .................... |
| V | 4366 hCoV-19/Japan/PG-0903/2020\|EPI_ISL_684513\|2020-03 | .................... | .................... | .................... | .................... | .................... | .................... |
| V | 4367 hCoV-19/Japan/PG-0904/2020\|EPI_ISL_684514\|2020-03 | .................... | .................... | .................... | .................... | .................... | .................... |
| V | 4368 hCoV-19/Japan/PG-0905/2020\|EPI_ISL_684515\|2020-04 | .................... | .................... | .................... | .................... | .................... | .................... |
| V | 4369 hCoV-19/Japan/PG-0906/2020\|EPI_ISL_684516\|2020-04 | .................... | .................... | .................... | .................... | .................... | .................... |
| V | 4370 hCoV-19/Japan/PG-0907/2020\|EPI_ISL_684517\|2020-03 | .................... | .................... | .................... | .................... | .................... | .................... |
| V | 4371 hCoV-19/Japan/PG-0908/2020\|EPI_ISL_684518\|2020-04 | .................... | .................... | .................... | .................... | .................... | .................... |
| V | 4372 hCoV-19/Japan/PG-0909/2020\|EPI_ISL_684519\|2020-03 | .................... | .................... | .................... | .................... | .................... | .................... |
| V | 4373 hCoV-19/Japan/PG-0910/2020\|EPI_ISL_684520\|2020-03 | .................... | .................... | .................... | .................... | .................... | .................... |
| V | 4374 hCoV-19/Japan/PG-0911/2020\|EPI_ISL_684521\|2020-03 | .................... | .................... | .................... | .................... | .................... | .................... |
| V | 4375 hCoV-19/Japan/PG-0912/2020\|EPI_ISL_684522\|2020-03 | .................... | .................... | .................... | .................... | .................... | .................... |
| V | 4376 hCoV-19/Japan/PG-0913/2020\|EPI_ISL_684523\|2020-04 | .................... | .................... | .................... | .................... | .................... | .................... |
| V | 4377 hCoV-19/Japan/PG-0914/2020\|EPI_ISL_684524\|2020-04 | .................... | .................... | .................... | .................... | .................... | .................... |
| V | 4378 hCoV-19/Japan/PG-0915/2020\|EPI_ISL_684525\|2020-04 | .................... | .................... | .................... | .................... | .................... | .................... |
| V | 4379 hCoV-19/Japan/PG-0916/2020\|EPI_ISL_684526\|2020-04 | .................... | .................... | .................... | .................... | .................... | .................... |
| V | 4380 hCoV-19/Japan/PG-0917/2020\|EPI_ISL_684527\|2020-04 | .................... | .................... | .................... | .................... | .................... | .................... |
| V | 4381 hCoV-19/Japan/PG-0919/2020\|EPI_ISL_684529\|2020-04 | .................... | .................... | .................... | .................... | .................... | .................... |
| V | 4382 hCoV-19/Japan/PG-0920/2020\|EPI_ISL_684530\|2020-04 | .................... | .................... | .................... | .................... | .................... | .................... |
| V | 4383 hCoV-19/Japan/PG-0921/2020\|EPI_ISL_684531\|2020-04 | .................... | .................... | .................... | .................... | .................... | .................... |
| V | 4384 hCoV-19/Japan/PG-0922/2020\|EPI_ISL_684532\|2020-04 | .................... | .................... | .................... | .................... | .................... | .................... |
| V | 4385 hCoV-19/Japan/PG-0923/2020\|EPI_ISL_684533\|2020-04 | .................... | .................... | .................... | .................... | .................... | .................... |
| V | 4386 hCoV-19/Japan/PG-0924/2020\|EPI_ISL_684534\|2020-04 | .................... | .................... | .................... | .................... | .................... | .................... |
| V | 4387 hCoV-19/Japan/PG-0925/2020\|EPI_ISL_684535\|2020-04 | .................... | .................... | .................... | .................... | .................... | .................... |
| V | 4388 hCoV-19/Japan/PG-0926/2020\|EPI_ISL_684536\|2020-04 | .................... | .................... | .................... | .................... | .................... | .................... |
| V | 4389 hCoV-19/Japan/PG-0927/2020\|EPI_ISL_684537\|2020-04 | .................... | .................... | .................... | .................... | .................... | .................... |
| V | 4390 hCoV-19/Japan/PG-0928/2020\|EPI_ISL_684538\|2020-04 | .................... | .................... | .................... | .................... | .................... | .................... |
| V | 4391 hCoV-19/Japan/PG-0929/2020\|EPI_ISL_684539\|2020-04 | .................... | .................... | .................... | .................... | .................... | .................... |
| V | 4392 hCoV-19/Japan/PG-0931/2020\|EPI_ISL_684541\|2020-04 | .................... | .................... | .................... | .................... | .................... | .................... |
| V | 4393 hCoV-19/Japan/PG-0932/2020\|EPI_ISL_684542\|2020-04 | .................... | .................... | .................... | .................... | .................... | .................... |
| V | 4394 hCoV-19/Japan/PG-0944/2020\|EPI_ISL_684554\|2020-04 | .................... | .................... | .................... | .................... | .................... | .................... |
| V | 4395 hCoV-19/Japan/PG-0945/2020\|EPI_ISL_684555\|2020-04 | .................... | .................... | .................... | .................... | .................... | .................... |
| V | 4396 hCoV-19/Japan/PG-0946/2020\|EPI_ISL_684556\|2020-03 | .................... | .................... | .................... | .................... | .................... | .................... |
| V | 4397 hCoV-19/Japan/PG-0947/2020\|EPI_ISL_684557\|2020-04 | .................... | .................... | .................... | .................... | .................... | .................... |

| | | | | | | | | |
|---|---|---|---|---|---|---|---|---|
| V | 4398 | hCoV-19/Japan/PG-0948/2020\|EPI_ISL_684558\|2020-04 | .................... | .................... | ..................... | .................... | .................... | .................... |
| V | 4399 | hCoV-19/Japan/PG-0949/2020\|EPI_ISL_684559\|2020-04 | .................... | .................... | ..................... | .................... | .................... | .................... |
| V | 4400 | hCoV-19/Japan/PG-0950/2020\|EPI_ISL_684560\|2020-04 | .................... | .................... | ..................... | .................... | .................... | .................... |
| V | 4401 | hCoV-19/Japan/PG-0951/2020\|EPI_ISL_684561\|2020-04 | .................... | .................... | ..................... | .................... | .................... | .................... |
| V | 4402 | hCoV-19/Japan/PG-0952/2020\|EPI_ISL_684562\|2020-04 | .................... | .................... | ..................... | .................... | .................... | .................... |
| V | 4403 | hCoV-19/Japan/PG-0953/2020\|EPI_ISL_684563\|2020-04 | .................... | .................... | ..................... | .................... | .................... | .................... |
| V | 4404 | hCoV-19/Japan/PG-0954/2020\|EPI_ISL_684564\|2020-04 | .................... | .................... | ..................... | .................... | .................... | .................... |
| V | 4405 | hCoV-19/Japan/PG-0955/2020\|EPI_ISL_684565\|2020-04 | .................... | .................... | ..................... | .................... | .................... | .................... |
| V | 4406 | hCoV-19/Japan/PG-0956/2020\|EPI_ISL_684566\|2020-04 | .................... | .................... | ..................... | .................... | .................... | .................... |
| V | 4407 | hCoV-19/Japan/PG-0957/2020\|EPI_ISL_684567\|2020-04 | .................... | .................... | ..................... | .................... | .................... | .................... |
| V | 4408 | hCoV-19/Japan/PG-0958/2020\|EPI_ISL_684568\|2020-04 | .................... | .................... | ..................... | .................... | .................... | .................... |
| V | 4409 | hCoV-19/Japan/PG-0959/2020\|EPI_ISL_684569\|2020-04 | .................... | .................... | ..................... | .................... | .................... | .................... |
| V | 4410 | hCoV-19/Japan/PG-0960/2020\|EPI_ISL_684570\|2020-04 | .................... | .................... | ..................... | .................... | .................... | .................... |
| V | 4411 | hCoV-19/Japan/PG-0961/2020\|EPI_ISL_684571\|2020-04 | .................... | .................... | ..................... | .................... | .................... | .................... |
| V | 4412 | hCoV-19/Japan/PG-0962/2020\|EPI_ISL_684572\|2020-04 | .................... | .................... | ..................... | .................... | .................... | .................... |
| V | 4413 | hCoV-19/Japan/PG-0963/2020\|EPI_ISL_684573\|2020-04 | .................... | .................... | ..................... | .................... | .................... | .................... |
| V | 4414 | hCoV-19/Japan/PG-0964/2020\|EPI_ISL_684574\|2020-04 | .................... | .................... | ..................... | .................... | .................... | .................... |
| V | 4415 | hCoV-19/Japan/PG-0965/2020\|EPI_ISL_684575\|2020-04 | .................... | .................... | ..................... | .................... | .................... | .................... |
| V | 4416 | hCoV-19/Japan/PG-0966/2020\|EPI_ISL_684576\|2020-04 | .................... | .................... | ..................... | .................... | .................... | .................... |
| V | 4417 | hCoV-19/Japan/PG-0968/2020\|EPI_ISL_684578\|2020-04 | .................... | .................... | ..................... | .................... | .................... | .................... |
| V | 4418 | hCoV-19/Japan/PG-0969/2020\|EPI_ISL_684579\|2020-04 | .................... | .................... | ..................... | .................... | .................... | .................... |
| V | 4419 | hCoV-19/Japan/PG-0970/2020\|EPI_ISL_684580\|2020-04 | .................... | .................... | ..................... | .................... | .................... | .................... |
| V | 4420 | hCoV-19/Japan/PG-0971/2020\|EPI_ISL_684581\|2020-04 | .................... | .................... | ..................... | .................... | .................... | .................... |
| V | 4421 | hCoV-19/Japan/PG-0972/2020\|EPI_ISL_684582\|2020-04 | .................... | .................... | ..................... | .................... | .................... | .................... |
| V | 4422 | hCoV-19/Japan/PG-0973/2020\|EPI_ISL_684583\|2020-04 | .................... | .................... | ..................... | .................... | .................... | .................... |
| V | 4423 | hCoV-19/Japan/PG-0974/2020\|EPI_ISL_684584\|2020-04 | .................... | .................... | ..................... | .................... | .................... | .................... |
| V | 4424 | hCoV-19/Japan/PG-0975/2020\|EPI_ISL_684585\|2020-04 | .................... | .................... | ..................... | .................... | .................... | .................... |
| V | 4425 | hCoV-19/Japan/PG-0976/2020\|EPI_ISL_684586\|2020-04 | .................... | .................... | ..................... | .................... | .................... | .................... |
| V | 4426 | hCoV-19/Japan/PG-0977/2020\|EPI_ISL_684587\|2020-04 | .................... | .................... | ..................... | .................... | .................... | .................... |
| V | 4427 | hCoV-19/Japan/PG-0978/2020\|EPI_ISL_684588\|2020-04 | .................... | .................... | ..................... | .................... | .................... | .................... |
| V | 4428 | hCoV-19/Japan/PG-0979/2020\|EPI_ISL_684589\|2020-04 | .................... | .................... | ..................... | .................... | .................... | .................... |
| V | 4429 | hCoV-19/Japan/PG-0980/2020\|EPI_ISL_684590\|2020-04 | .................... | .................... | ..................... | .................... | .................... | .................... |
| V | 4430 | hCoV-19/Japan/PG-0981/2020\|EPI_ISL_684591\|2020-04 | .................... | .................... | ..................... | .................... | .................... | .................... |
| V | 4431 | hCoV-19/Japan/PG-0982/2020\|EPI_ISL_684592\|2020-04 | .................... | .................... | ..................... | .................... | .................... | .................... |
| V | 4432 | hCoV-19/Japan/PG-0983/2020\|EPI_ISL_684593\|2020-04 | .................... | .................... | ..................... | .................... | .................... | .................... |
| V | 4433 | hCoV-19/Japan/PG-0984/2020\|EPI_ISL_684594\|2020-04 | .................... | .................... | ..................... | .................... | .................... | .................... |
| V | 4434 | hCoV-19/Japan/PG-0985/2020\|EPI_ISL_684595\|2020-04 | .................... | .................... | ..................... | .................... | .................... | .................... |
| V | 4435 | hCoV-19/Japan/PG-0986/2020\|EPI_ISL_684596\|2020-04 | .................... | .................... | ..................... | .................... | .................... | .................... |
| V | 4436 | hCoV-19/Japan/PG-0987/2020\|EPI_ISL_684597\|2020-04 | .................... | .................... | ..................... | .................... | .................... | .................... |
| V | 4437 | hCoV-19/Japan/PG-0988/2020\|EPI_ISL_684598\|2020-04 | .................... | .................... | ..................... | .................... | .................... | .................... |
| V | 4438 | hCoV-19/Japan/PG-0989/2020\|EPI_ISL_684599\|2020-04 | .................... | .................... | ..................... | .................... | .................... | .................... |
| V | 4439 | hCoV-19/Japan/PG-2195/2020\|EPI_ISL_685800\|2020-04 | .................... | .................... | ..................... | .................... | .................... | .................... |
| V | 4440 | hCoV-19/Japan/PG-2196/2020\|EPI_ISL_685801\|2020-04 | .................... | .................... | ..................... | .................... | .................... | .................... |
| V | 4441 | hCoV-19/Japan/PG-2197/2020\|EPI_ISL_685802\|2020-04 | .................... | .................... | ..................... | .................... | .................... | .................... |
| V | 4442 | hCoV-19/Japan/PG-2198/2020\|EPI_ISL_685803\|2020-04 | .................... | .................... | ..................... | .................... | .................... | .................... |
| V | 4443 | hCoV-19/Japan/PG-2199/2020\|EPI_ISL_685804\|2020-04 | .................... | .................... | ..................... | .................... | .................... | .................... |
| V | 4444 | hCoV-19/Japan/PG-2200/2020\|EPI_ISL_685805\|2020-04 | .................... | .................... | ..................... | .................... | .................... | .................... |
| V | 4445 | hCoV-19/Japan/PG-2201/2020\|EPI_ISL_685806\|2020-04 | .................... | .................... | ..................... | .................... | .................... | .................... |
| V | 4446 | hCoV-19/Japan/PG-2202/2020\|EPI_ISL_685807\|2020-04 | .................... | .................... | ..................... | .................... | .................... | .................... |
| V | 4447 | hCoV-19/Japan/PG-2203/2020\|EPI_ISL_685808\|2020-04 | .................... | .................... | ..................... | .................... | .................... | .................... |
| V | 4448 | hCoV-19/Japan/PG-2204/2020\|EPI_ISL_685809\|2020-04 | .................... | .................... | ..................... | .................... | .................... | .................... |
| V | 4449 | hCoV-19/Japan/PG-2205/2020\|EPI_ISL_685810\|2020-04 | .................... | .................... | ..................... | .................... | .................... | .................... |
| V | 4450 | hCoV-19/Japan/PG-2206/2020\|EPI_ISL_685811\|2020-04 | .................... | .................... | ..................... | .................... | .................... | .................... |
| V | 4451 | hCoV-19/Japan/PG-2207/2020\|EPI_ISL_685812\|2020-04 | .................... | .................... | ..................... | .................... | .................... | .................... |
| V | 4452 | hCoV-19/Japan/PG-2208/2020\|EPI_ISL_685813\|2020-04 | .................... | .................... | ..................... | .................... | .................... | .................... |
| V | 4453 | hCoV-19/Japan/PG-2209/2020\|EPI_ISL_685814\|2020-04 | .................... | .................... | ..................... | .................... | .................... | .................... |
| V | 4454 | hCoV-19/Japan/PG-2210/2020\|EPI_ISL_685815\|2020-04 | .................... | .................... | ..................... | .................... | .................... | .................... |
| V | 4455 | hCoV-19/Japan/PG-2211/2020\|EPI_ISL_685816\|2020-04 | .................... | .................... | ..................... | .................... | .................... | .................... |
| V | 4456 | hCoV-19/Japan/PG-2212/2020\|EPI_ISL_685817\|2020-04 | .................... | .................... | ..................... | .................... | .................... | .................... |
| V | 4457 | hCoV-19/Japan/PG-2213/2020\|EPI_ISL_685818\|2020-04 | .................... | .................... | ..................... | .................... | .................... | .................... |
| V | 4458 | hCoV-19/Japan/PG-2214/2020\|EPI_ISL_685819\|2020-04 | .................... | .................... | ..................... | .................... | .................... | .................... |
| V | 4459 | hCoV-19/Japan/PG-2215/2020\|EPI_ISL_685820\|2020-04 | .................... | .................... | ..................... | .................... | .................... | .................... |
| V | 4460 | hCoV-19/Japan/PG-2216/2020\|EPI_ISL_685821\|2020-04 | .................... | .................... | ..................... | .................... | .................... | .................... |
| V | 4461 | hCoV-19/Japan/PG-2217/2020\|EPI_ISL_685822\|2020-04 | .................... | .................... | ..................... | .................... | .................... | .................... |
| V | 4462 | hCoV-19/Japan/PG-2218/2020\|EPI_ISL_685823\|2020-04 | .................... | .................... | ..................... | .................... | .................... | .................... |
| V | 4463 | hCoV-19/Japan/PG-2219/2020\|EPI_ISL_685824\|2020-04 | .................... | .................... | ..................... | .................... | .................... | .................... |
| V | 4464 | hCoV-19/Japan/PG-2220/2020\|EPI_ISL_685825\|2020-04 | .................... | .................... | ..................... | .................... | .................... | .................... |
| V | 4465 | hCoV-19/Japan/PG-2221/2020\|EPI_ISL_685826\|2020-04 | .................... | .................... | ..................... | .................... | .................... | .................... |
| V | 4466 | hCoV-19/Japan/PG-2222/2020\|EPI_ISL_685827\|2020-04 | .................... | .................... | ..................... | .................... | .................... | .................... |
| V | 4467 | hCoV-19/Japan/PG-2223/2020\|EPI_ISL_685828\|2020-04 | .................... | .................... | ..................... | .................... | .................... | .................... |
| V | 4468 | hCoV-19/Japan/PG-2224/2020\|EPI_ISL_685829\|2020-04 | .................... | .................... | ..................... | .................... | .................... | .................... |
| V | 4469 | hCoV-19/Japan/PG-2225/2020\|EPI_ISL_685830\|2020-04 | .................... | .................... | ..................... | .................... | .................... | .................... |
| V | 4470 | hCoV-19/Japan/PG-2226/2020\|EPI_ISL_685831\|2020-04 | .................... | .................... | ..................... | .................... | .................... | .................... |
| V | 4471 | hCoV-19/Japan/PG-2227/2020\|EPI_ISL_685832\|2020-04 | .................... | .................... | ..................... | .................... | .................... | .................... |
| V | 4472 | hCoV-19/Japan/PG-2228/2020\|EPI_ISL_685833\|2020-04 | .................... | .................... | ..................... | .................... | .................... | .................... |
| V | 4473 | hCoV-19/Japan/PG-2229/2020\|EPI_ISL_685834\|2020-04 | .................... | .................... | ..................... | .................... | .................... | .................... |
| V | 4474 | hCoV-19/Japan/PG-2230/2020\|EPI_ISL_685835\|2020-04 | .................... | .................... | ..................... | .................... | .................... | .................... |
| V | 4475 | hCoV-19/Japan/PG-2231/2020\|EPI_ISL_685836\|2020-04 | .................... | .................... | ..................... | .................... | .................... | .................... |
| V | 4476 | hCoV-19/Japan/PG-2232/2020\|EPI_ISL_685837\|2020-04 | .................... | .................... | ..................... | .................... | .................... | .................... |
| V | 4477 | hCoV-19/Japan/PG-2233/2020\|EPI_ISL_685838\|2020-04 | .................... | .................... | ..................... | .................... | .................... | .................... |

| | | | | | | | | |
|---|---|---|---|---|---|---|---|---|
| V | 4478 | hCoV-19/Japan/PG-2234/2020\|EPI_ISL_685839\|2020-04 | ···················· | ···················· | ····················· | ···················· | ···················· | ···················· |
| V | 4479 | hCoV-19/Japan/PG-2235/2020\|EPI_ISL_685840\|2020-04 | ···················· | ···················· | ····················· | ···················· | ···················· | ···················· |
| V | 4480 | hCoV-19/Japan/PG-2236/2020\|EPI_ISL_685841\|2020-04 | ···················· | ···················· | ····················· | ···················· | ···················· | ···················· |
| V | 4481 | hCoV-19/Japan/PG-2237/2020\|EPI_ISL_685842\|2020-04 | ···················· | ···················· | ····················· | ···················· | ···················· | ···················· |
| V | 4482 | hCoV-19/Japan/PG-2238/2020\|EPI_ISL_685843\|2020-04 | ···················· | ···················· | ····················· | ···················· | ···················· | ···················· |
| V | 4483 | hCoV-19/Japan/PG-2239/2020\|EPI_ISL_685844\|2020-04 | ···················· | ···················· | ····················· | ···················· | ···················· | ···················· |
| V | 4484 | hCoV-19/Japan/PG-2240/2020\|EPI_ISL_685845\|2020-04 | ···················· | ···················· | ····················· | ···················· | ···················· | ···················· |
| V | 4485 | hCoV-19/Japan/PG-2241/2020\|EPI_ISL_685846\|2020-04 | ···················· | ···················· | ····················· | ···················· | ···················· | ···················· |
| V | 4486 | hCoV-19/Japan/PG-2242/2020\|EPI_ISL_685847\|2020-04 | ···················· | ···················· | ····················· | ···················· | ···················· | ···················· |
| V | 4487 | hCoV-19/Japan/PG-2243/2020\|EPI_ISL_685848\|2020-04 | ···················· | ···················· | ····················· | ···················· | ···················· | ···················· |
| V | 4488 | hCoV-19/Japan/PG-2244/2020\|EPI_ISL_685849\|2020-04 | ···················· | ···················· | ····················· | ···················· | ···················· | ···················· |
| V | 4489 | hCoV-19/Japan/PG-2245/2020\|EPI_ISL_685850\|2020-04 | ···················· | ···················· | ····················· | ···················· | ···················· | ···················· |
| V | 4490 | hCoV-19/Japan/PG-2246/2020\|EPI_ISL_685851\|2020-04 | ···················· | ···················· | ····················· | ···················· | ···················· | ···················· |
| V | 4491 | hCoV-19/Japan/PG-2247/2020\|EPI_ISL_685852\|2020-04 | ···················· | ···················· | ····················· | ···················· | ···················· | ···················· |
| V | 4492 | hCoV-19/Japan/PG-2248/2020\|EPI_ISL_685853\|2020-04 | ···················· | ···················· | ····················· | ···················· | ···················· | ···················· |
| V | 4493 | hCoV-19/Japan/PG-2249/2020\|EPI_ISL_685854\|2020-04 | ···················· | ···················· | ····················· | ···················· | ···················· | ···················· |
| V | 4494 | hCoV-19/Japan/PG-2250/2020\|EPI_ISL_685855\|2020-04 | ···················· | ···················· | ····················· | ···················· | ···················· | ···················· |
| V | 4495 | hCoV-19/Japan/PG-2251/2020\|EPI_ISL_685856\|2020-04 | ···················· | ···················· | ····················· | ···················· | ···················· | ···················· |
| V | 4496 | hCoV-19/Japan/PG-2252/2020\|EPI_ISL_685857\|2020-04 | ···················· | ···················· | ····················· | ···················· | ···················· | ···················· |
| V | 4497 | hCoV-19/Japan/PG-2253/2020\|EPI_ISL_685858\|2020-04 | ···················· | ···················· | ····················· | ···················· | ···················· | ···················· |
| V | 4498 | hCoV-19/Japan/PG-2254/2020\|EPI_ISL_685859\|2020-04 | ···················· | ···················· | ····················· | ···················· | ···················· | ···················· |
| V | 4499 | hCoV-19/Japan/PG-2255/2020\|EPI_ISL_685860\|2020-04 | ···················· | ···················· | ····················· | ···················· | ···················· | ···················· |
| V | 4500 | hCoV-19/Japan/PG-2256/2020\|EPI_ISL_685861\|2020-04 | ···················· | ···················· | ····················· | ···················· | ···················· | ···················· |
| V | 4501 | hCoV-19/Japan/PG-2257/2020\|EPI_ISL_685862\|2020-04 | ···················· | ···················· | ····················· | ···················· | ···················· | ···················· |
| V | 4502 | hCoV-19/Japan/PG-2258/2020\|EPI_ISL_685863\|2020-04 | ···················· | ···················· | ····················· | ···················· | ···················· | ···················· |
| V | 4503 | hCoV-19/Japan/PG-2259/2020\|EPI_ISL_685864\|2020-04 | ···················· | ···················· | ····················· | ···················· | ···················· | ···················· |
| V | 4504 | hCoV-19/Japan/PG-2260/2020\|EPI_ISL_685865\|2020-04 | ···················· | ···················· | ····················· | ···················· | ···················· | ···················· |
| V | 4505 | hCoV-19/Japan/PG-2261/2020\|EPI_ISL_685866\|2020-04 | ···················· | ···················· | ····················· | ···················· | ···················· | ···················· |
| V | 4506 | hCoV-19/Japan/PG-2262/2020\|EPI_ISL_685867\|2020-04 | ···················· | ···················· | ····················· | ···················· | ···················· | ···················· |
| V | 4507 | hCoV-19/Japan/PG-2263/2020\|EPI_ISL_685868\|2020-04 | ···················· | ···················· | ····················· | ···················· | ···················· | ···················· |
| V | 4508 | hCoV-19/Japan/PG-2264/2020\|EPI_ISL_685869\|2020-04 | ···················· | ···················· | ····················· | ···················· | ···················· | ···················· |
| V | 4509 | hCoV-19/Japan/PG-2266/2020\|EPI_ISL_685871\|2020-04 | ···················· | ···················· | ····················· | ···················· | ···················· | ···················· |
| V | 4510 | hCoV-19/Japan/PG-2280/2020\|EPI_ISL_685885\|2020-04 | ···················· | ····G··············· | ····················· | ···················· | ···················· | ···················· |
| V | 4511 | hCoV-19/Japan/PG-2281/2020\|EPI_ISL_685886\|2020-04 | ···················· | ···················· | ····················· | ···················· | ···················· | ···················· |
| V | 4512 | hCoV-19/Japan/PG-2282/2020\|EPI_ISL_685887\|2020-04 | ···················· | ···················· | ····················· | ···················· | ···················· | ···················· |
| V | 4513 | hCoV-19/Japan/PG-2283/2020\|EPI_ISL_685888\|2020-04 | ···················· | ···················· | ····················· | ···················· | ···················· | ···················· |
| V | 4514 | hCoV-19/Japan/PG-2284/2020\|EPI_ISL_685889\|2020-04 | ···················· | ···················· | ····················· | ···················· | ···················· | ···················· |
| V | 4515 | hCoV-19/Japan/PG-2285/2020\|EPI_ISL_685890\|2020-04 | ···················· | ···················· | ····················· | ···················· | ···················· | ···················· |
| V | 4516 | hCoV-19/Japan/PG-2286/2020\|EPI_ISL_685891\|2020-04 | ···················· | ···················· | ····················· | ···················· | ···················· | ···················· |
| V | 4517 | hCoV-19/Japan/PG-2287/2020\|EPI_ISL_685892\|2020-04 | ···················· | ···················· | ····················· | ···················· | ···················· | ···················· |
| V | 4518 | hCoV-19/Japan/PG-2288/2020\|EPI_ISL_685893\|2020-04 | ···················· | ···················· | ····················· | ···················· | ···················· | ···················· |
| V | 4519 | hCoV-19/Japan/PG-2289/2020\|EPI_ISL_685894\|2020-04 | ···················· | ···················· | ····················· | ···················· | ···················· | ···················· |
| V | 4520 | hCoV-19/Japan/PG-2290/2020\|EPI_ISL_685895\|2020-04 | ···················· | ···················· | ····················· | ···················· | ···················· | ···················· |
| V | 4521 | hCoV-19/Japan/PG-2291/2020\|EPI_ISL_685896\|2020-04 | ···················· | ···················· | ····················· | ···················· | ···················· | ···················· |
| V | 4522 | hCoV-19/Japan/PG-2292/2020\|EPI_ISL_685897\|2020-04 | ···················· | ···················· | ····················· | ···················· | ···················· | ···················· |
| V | 4523 | hCoV-19/Japan/PG-2293/2020\|EPI_ISL_685898\|2020-04 | ···················· | ···················· | ····················· | ···················· | ···················· | ···················· |
| V | 4524 | hCoV-19/Japan/PG-2294/2020\|EPI_ISL_685899\|2020-04 | ···················· | ···················· | ····················· | ···················· | ···················· | ···················· |
| V | 4525 | hCoV-19/Japan/PG-6975/2020\|EPI_ISL_688080\|2020-07 | ···················· | ···················· | ····················· | ···················· | ···················· | ···················· |
| V | 4526 | hCoV-19/Japan/PG-6976/2020\|EPI_ISL_688081\|2020-07 | ···················· | ···················· | ····················· | ···················· | ···················· | ···················· |
| V | 4527 | hCoV-19/Japan/PG-6977/2020\|EPI_ISL_688082\|2020-07 | ···················· | ···················· | ····················· | ···················· | ···················· | ···················· |
| V | 4528 | hCoV-19/Japan/PG-6978/2020\|EPI_ISL_688083\|2020-07 | ···················· | ···················· | ····················· | ···················· | ···················· | ···················· |
| V | 4529 | hCoV-19/Japan/PG-6979/2020\|EPI_ISL_688084\|2020-04 | ···················· | ···················· | ····················· | ···················· | ···················· | ···················· |
| V | 4530 | hCoV-19/Japan/PG-6980/2020\|EPI_ISL_688085\|2020-04 | ···················· | ···················· | ····················· | ···················· | ···················· | ···················· |
| V | 4531 | hCoV-19/Japan/PG-6981/2020\|EPI_ISL_688086\|2020-07 | ···················· | ···················· | ····················· | ···················· | ···················· | ···················· |
| V | 4532 | hCoV-19/Japan/PG-6982/2020\|EPI_ISL_688087\|2020-07 | ···················· | ···················· | ····················· | ···················· | ···················· | ···················· |
| V | 4533 | hCoV-19/Japan/PG-6983/2020\|EPI_ISL_688088\|2020-07 | ···················· | ···················· | ····················· | ···················· | ···················· | ···················· |
| V | 4534 | hCoV-19/Japan/PG-6984/2020\|EPI_ISL_688089\|2020-07 | ···················· | ···················· | ····················· | ···················· | ···················· | ···················· |
| V | 4535 | hCoV-19/Japan/PG-6985/2020\|EPI_ISL_688090\|2020-07 | ···················· | ···················· | ····················· | ···················· | ···················· | ···················· |
| V | 4536 | hCoV-19/Japan/PG-6986/2020\|EPI_ISL_688091\|2020-07 | ···················· | ···················· | ····················· | ···················· | ···················· | ···················· |
| V | 4537 | hCoV-19/Japan/PG-6987/2020\|EPI_ISL_688092\|2020-07 | ···················· | ···················· | ····················· | ···················· | ···················· | ···················· |
| V | 4538 | hCoV-19/Japan/PG-6988/2020\|EPI_ISL_688093\|2020-07 | ···················· | ···················· | ····················· | ···················· | ···················· | ···················· |
| V | 4539 | hCoV-19/Japan/PG-6989/2020\|EPI_ISL_688094\|2020-08 | ···················· | ···················· | ····················· | ···················· | ···················· | ···················· |
| V | 4540 | hCoV-19/Japan/PG-6990/2020\|EPI_ISL_688095\|2020-08 | ···················· | ···················· | ····················· | ···················· | ···················· | ···················· |
| V | 4541 | hCoV-19/Japan/PG-6991/2020\|EPI_ISL_688096\|2020-08 | ···················· | ···················· | ····················· | ···················· | ···················· | ···················· |
| V | 4542 | hCoV-19/Japan/PG-6992/2020\|EPI_ISL_688097\|2020-08 | ···················· | ···················· | ····················· | ···················· | ···················· | ···················· |
| V | 4543 | hCoV-19/Japan/PG-6993/2020\|EPI_ISL_688098\|2020-08 | ···················· | ···················· | ····················· | ···················· | ···················· | ···················· |
| V | 4544 | hCoV-19/Japan/PG-6994/2020\|EPI_ISL_688099\|2020-08 | ···················· | ···················· | ····················· | ···················· | ···················· | ···················· |
| V | 4545 | hCoV-19/Japan/PG-6192/2020\|EPI_ISL_689304\|2020-08 | ···················· | ···················· | ····················· | ···················· | ···················· | ···················· |
| V | 4546 | hCoV-19/Japan/PG-6193/2020\|EPI_ISL_689305\|2020-08 | ···················· | ···················· | ····················· | ···················· | ···················· | ···················· |
| V | 4547 | hCoV-19/Japan/PG-6194/2020\|EPI_ISL_689306\|2020-08 | ···················· | ···················· | ····················· | ···················· | ···················· | ···················· |
| V | 4548 | hCoV-19/Japan/PG-6195/2020\|EPI_ISL_689307\|2020-08 | ···················· | ···················· | ····················· | ···················· | ···················· | ···················· |
| V | 4549 | hCoV-19/Japan/PG-6196/2020\|EPI_ISL_689308\|2020-08 | ···················· | ···················· | ····················· | ···················· | ···················· | ···················· |
| V | 4550 | hCoV-19/Japan/PG-6197/2020\|EPI_ISL_689309\|2020-08 | ···················· | ···················· | ····················· | ···················· | ···················· | ···················· |
| V | 4551 | hCoV-19/Japan/PG-6198/2020\|EPI_ISL_689310\|2020-08 | ···················· | ···················· | ····················· | ···················· | ···················· | ···················· |
| V | 4552 | hCoV-19/Japan/PG-6199/2020\|EPI_ISL_689311\|2020-08 | ···················· | ···················· | ····················· | ···················· | ···················· | ···················· |
| V | 4553 | hCoV-19/Japan/PG-6200/2020\|EPI_ISL_689312\|2020-08 | ···················· | ···················· | ····················· | ···················· | ···················· | ···················· |
| V | 4554 | hCoV-19/Japan/PG-6201/2020\|EPI_ISL_689313\|2020-08 | ···················· | ···················· | ····················· | ···················· | ···················· | ···················· |
| V | 4555 | hCoV-19/Japan/PG-6202/2020\|EPI_ISL_689314\|2020-08 | ···················· | ···················· | ····················· | ···················· | ···················· | ···················· |
| V | 4556 | hCoV-19/Japan/PG-6203/2020\|EPI_ISL_689315\|2020-08 | ···················· | ···················· | ····················· | ···················· | ···················· | ···················· |
| V | 4557 | hCoV-19/Japan/PG-6204/2020\|EPI_ISL_689316\|2020-08 | ···················· | ···················· | ····················· | ···················· | ···················· | ···················· |

| | | | | | | | | |
|---|---|---|---|---|---|---|---|---|
| V | 4558 | hCoV-19/Japan/PG-6205/2020\|EPI_ISL_689317\|2020-08 | .................... | .................... | .................... | .................... | .................... | .................... |
| V | 4559 | hCoV-19/Japan/PG-6206/2020\|EPI_ISL_689318\|2020-08 | .................... | .................... | .................... | .................... | .................... | .................... |
| V | 4560 | hCoV-19/Japan/PG-6207/2020\|EPI_ISL_689319\|2020-08 | .................... | .................... | .................... | .................... | .................... | .................... |
| V | 4561 | hCoV-19/Japan/PG-6208/2020\|EPI_ISL_689320\|2020-08 | .................... | .................... | .................... | .................... | .................... | .................... |
| V | 4562 | hCoV-19/Japan/PG-6209/2020\|EPI_ISL_689321\|2020-08 | .................... | .................... | .................... | .................... | .................... | .................... |
| V | 4563 | hCoV-19/Japan/PG-6210/2020\|EPI_ISL_689322\|2020-08 | .................... | .................... | .................... | .................... | .................... | .................... |
| V | 4564 | hCoV-19/Japan/PG-6211/2020\|EPI_ISL_689323\|2020-08 | .................... | .................... | .................... | .................... | .................... | .................... |
| V | 4565 | hCoV-19/Japan/PG-6212/2020\|EPI_ISL_689324\|2020-08 | .................... | .................... | .................... | .................... | .................... | .................... |
| V | 4566 | hCoV-19/Japan/PG-6213/2020\|EPI_ISL_689325\|2020-08 | .................... | .................... | .................... | .................... | .................... | .................... |
| V | 4567 | hCoV-19/Japan/PG-6214/2020\|EPI_ISL_689326\|2020-08 | .................... | .................... | .................... | .................... | .................... | .................... |
| V | 4568 | hCoV-19/Japan/PG-6215/2020\|EPI_ISL_689327\|2020-08 | .................... | .................... | .................... | .................... | .................... | .................... |
| V | 4569 | hCoV-19/Japan/PG-5207/2020\|EPI_ISL_690310\|2020-08 | .................... | .................... | .................... | .................... | .................... | .................... |
| V | 4570 | hCoV-19/Japan/PG-5208/2020\|EPI_ISL_690311\|2020-08 | .................... | .................... | .................... | .................... | .................... | .................... |
| V | 4571 | hCoV-19/Japan/PG-5209/2020\|EPI_ISL_690312\|2020-08 | .................... | .................... | .................... | .................... | .................... | .................... |
| V | 4572 | hCoV-19/Japan/PG-5210/2020\|EPI_ISL_690313\|2020-08 | .................... | .................... | .................... | .................... | .................... | .................... |
| V | 4573 | hCoV-19/Japan/PG-5211/2020\|EPI_ISL_690314\|2020-08 | .................... | .................... | .................... | .................... | .................... | .................... |
| V | 4574 | hCoV-19/Japan/PG-5212/2020\|EPI_ISL_690315\|2020-08 | .................... | .................... | .................... | .................... | .................... | .................... |
| V | 4575 | hCoV-19/Japan/PG-5213/2020\|EPI_ISL_690316\|2020-08 | .................... | .................... | .................... | .................... | .................... | .................... |
| V | 4576 | hCoV-19/Japan/PG-5214/2020\|EPI_ISL_690317\|2020-08 | .................... | .................... | .................... | .................... | .................... | .................... |
| V | 4577 | hCoV-19/Japan/PG-5215/2020\|EPI_ISL_690318\|2020-08 | .................... | .................... | .................... | .................... | .................... | .................... |
| V | 4578 | hCoV-19/Japan/PG-5216/2020\|EPI_ISL_690319\|2020-08 | .................... | .................... | .................... | .................... | .................... | .................... |
| V | 4579 | hCoV-19/Japan/PG-5217/2020\|EPI_ISL_690320\|2020-08 | .................... | .................... | .................... | .................... | .................... | .................... |
| V | 4580 | hCoV-19/Japan/PG-5218/2020\|EPI_ISL_690321\|2020-08 | .................... | .................... | .................... | .................... | .................... | .................... |
| V | 4581 | hCoV-19/Japan/PG-5219/2020\|EPI_ISL_690322\|2020-08 | .................... | .................... | .................... | .................... | .................... | .................... |
| V | 4582 | hCoV-19/Japan/PG-5220/2020\|EPI_ISL_690323\|2020-08 | .................... | .................... | .................... | .................... | .................... | .................... |
| V | 4583 | hCoV-19/Japan/PG-5221/2020\|EPI_ISL_690324\|2020-08 | .................... | .................... | .................... | .................... | .................... | .................... |
| V | 4584 | hCoV-19/Japan/PG-5222/2020\|EPI_ISL_690325\|2020-08 | .................... | .................... | .................... | .................... | .................... | .................... |
| V | 4585 | hCoV-19/Japan/PG-5223/2020\|EPI_ISL_690326\|2020-08 | .................... | .................... | .................... | .................... | .................... | .................... |
| V | 4586 | hCoV-19/Japan/PG-5224/2020\|EPI_ISL_690327\|2020-08 | .................... | .................... | .................... | .................... | .................... | .................... |
| V | 4587 | hCoV-19/Japan/PG-5225/2020\|EPI_ISL_690328\|2020-08 | .................... | .................... | .................... | .................... | .................... | .................... |
| V | 4588 | hCoV-19/Japan/PG-5226/2020\|EPI_ISL_690329\|2020-08 | .................... | .................... | .................... | .................... | .................... | .................... |
| V | 4589 | hCoV-19/Japan/PG-5234/2020\|EPI_ISL_690337\|2020-08 | .................... | .................... | .................... | .................... | .................... | .................... |
| V | 4590 | hCoV-19/Japan/PG-5235/2020\|EPI_ISL_690338\|2020-08 | .................... | .................... | .................... | .................... | .................... | .................... |
| V | 4591 | hCoV-19/Japan/PG-5292/2020\|EPI_ISL_690395\|2020-08 | .................... | .................... | .................... | .................... | .................... | .................... |
| V | 4592 | hCoV-19/Japan/PG-5293/2020\|EPI_ISL_690396\|2020-08 | .................... | .................... | .................... | .................... | .................... | .................... |
| V | 4593 | hCoV-19/Japan/PG-5294/2020\|EPI_ISL_690397\|2020-08 | .................... | .................... | .................... | .................... | .................... | .................... |
| V | 4594 | hCoV-19/Japan/PG-5295/2020\|EPI_ISL_690398\|2020-08 | .................... | .................... | .................... | .................... | .................... | .................... |
| V | 4595 | hCoV-19/Japan/PG-5296/2020\|EPI_ISL_690399\|2020-08 | .................... | .................... | .................... | .................... | .................... | .................... |
| V | 4596 | hCoV-19/USA/MA-MASPHL-00666/2020\|EPI_ISL_692970\|20 | .................... | .................... | .................... | .................... | .................... | .................... |
| V | 4597 | hCoV-19/USA/MA-MASPHL-00664/2020\|EPI_ISL_692972\|20 | .................... | .................... | .................... | .................... | .................... | .................... |
| V | 4598 | hCoV-19/USA/MA-MASPHL-00661/2020\|EPI_ISL_692974\|20 | .................... | .................... | .................... | .................... | .................... | .................... |
| V | 4599 | hCoV-19/USA/MA-MASPHL-00658/2020\|EPI_ISL_692975\|20 | .................... | .................... | .................... | .................... | .................... | .................... |
| V | 4600 | hCoV-19/USA/MA-MASPHL-00657/2020\|EPI_ISL_692976\|20 | .................... | .................... | .................... | .................... | .................... | .................... |
| V | 4601 | hCoV-19/USA/MA-MASPHL-00656/2020\|EPI_ISL_692977\|20 | .................... | .................... | .................... | .................... | .................... | .................... |
| V | 4602 | hCoV-19/USA/MA-MASPHL-00655/2020\|EPI_ISL_692978\|20 | .................... | .................... | .................... | .................... | .................... | .................... |
| V | 4603 | hCoV-19/USA/MA-MASPHL-00653/2020\|EPI_ISL_692979\|20 | .................... | .................... | .................... | .................... | .................... | .................... |
| V | 4604 | hCoV-19/USA/MA-MASPHL-00651/2020\|EPI_ISL_692981\|20 | .................... | .................... | .................... | .................... | .................... | .................... |
| V | 4605 | hCoV-19/USA/MA-MASPHL-00648/2020\|EPI_ISL_692984\|20 | .................... | .................... | .................... | .................... | .................... | .................... |
| V | 4606 | hCoV-19/USA/MA-MASPHL-00646/2020\|EPI_ISL_692985\|20 | .................... | .................... | .................... | .................... | .................... | .................... |
| V | 4607 | hCoV-19/USA/MA-MASPHL-00645/2020\|EPI_ISL_692986\|20 | .................... | .................... | .................... | .................... | .................... | .................... |
| V | 4608 | hCoV-19/USA/MA-MASPHL-00640/2020\|EPI_ISL_692989\|20 | .................... | .................... | .................... | .................... | .................... | .................... |
| V | 4609 | hCoV-19/Japan/PG-6913/2020\|EPI_ISL_688021\|2020-09 | .................... | .................... | .................... | .................... | .................... | .................... |
| V | 4610 | hCoV-19/Japan/PG-6915/2020\|EPI_ISL_688022\|2020-09 | .................... | .................... | .................... | .................... | .................... | .................... |
| V | 4611 | hCoV-19/Japan/PG-6916/2020\|EPI_ISL_688023\|2020-09 | .................... | .................... | .................... | .................... | .................... | .................... |
| V | 4612 | hCoV-19/Japan/PG-6917/2020\|EPI_ISL_688024\|2020-09 | .................... | .................... | .................... | .................... | .................... | .................... |
| V | 4613 | hCoV-19/Japan/PG-6918/2020\|EPI_ISL_688025\|2020-09 | .................... | .................... | .................... | .................... | .................... | .................... |
| V | 4614 | hCoV-19/Japan/PG-6919/2020\|EPI_ISL_688026\|2020-09 | .................... | .................... | .................... | .................... | .................... | .................... |
| V | 4615 | hCoV-19/Japan/PG-6920/2020\|EPI_ISL_688027\|2020-09 | .................... | .................... | .................... | .................... | .................... | .................... |
| V | 4616 | hCoV-19/Japan/PG-6921/2020\|EPI_ISL_688028\|2020-09 | .................... | .................... | .................... | .................... | .................... | .................... |
| V | 4617 | hCoV-19/Japan/PG-6922/2020\|EPI_ISL_688029\|2020-09 | .................... | .................... | .................... | .................... | .................... | .................... |
| V | 4618 | hCoV-19/Japan/PG-6923/2020\|EPI_ISL_688030\|2020-09 | .................... | .................... | .................... | .................... | .................... | .................... |
| V | 4619 | hCoV-19/Japan/PG-6924/2020\|EPI_ISL_688031\|2020-09 | .................... | .................... | .................... | .................... | .................... | .................... |
| V | 4620 | hCoV-19/Japan/PG-6926/2020\|EPI_ISL_688032\|2020-09 | .................... | .................... | .................... | .................... | .................... | .................... |
| V | 4621 | hCoV-19/Japan/PG-6927/2020\|EPI_ISL_688033\|2020-09 | .................... | .................... | .................... | .................... | .................... | .................... |
| V | 4622 | hCoV-19/Japan/PG-6928/2020\|EPI_ISL_688034\|2020-09 | .................... | .................... | .................... | .................... | .................... | .................... |
| V | 4623 | hCoV-19/Japan/PG-6929/2020\|EPI_ISL_688035\|2020-09 | .................... | .................... | .................... | .................... | .................... | .................... |
| V | 4624 | hCoV-19/Japan/PG-6930/2020\|EPI_ISL_688036\|2020-09 | .................... | .................... | .................... | .................... | .................... | .................... |
| V | 4625 | hCoV-19/Japan/PG-6931/2020\|EPI_ISL_688037\|2020-09 | .................... | .................... | .................... | .................... | .................... | .................... |
| V | 4626 | hCoV-19/Japan/PG-6932/2020\|EPI_ISL_688038\|2020-09 | .................... | .................... | .................... | .................... | .................... | .................... |
| V | 4627 | hCoV-19/Japan/PG-6933/2020\|EPI_ISL_688039\|2020-09 | .................... | .................... | .................... | .................... | .................... | .................... |
| V | 4628 | hCoV-19/Japan/PG-6934/2020\|EPI_ISL_688040\|2020-09 | .................... | .................... | .................... | .................... | .................... | .................... |
| V | 4629 | hCoV-19/Japan/PG-6935/2020\|EPI_ISL_688041\|2020-09 | .................... | .................... | .................... | .................... | .................... | .................... |
| V | 4630 | hCoV-19/Japan/PG-6936/2020\|EPI_ISL_688042\|2020-09 | .................... | .................... | .................... | .................... | .................... | .................... |
| V | 4631 | hCoV-19/Japan/PG-6937/2020\|EPI_ISL_688043\|2020-09 | .................... | .................... | .................... | .................... | .................... | .................... |
| V | 4632 | hCoV-19/Japan/PG-6938/2020\|EPI_ISL_688044\|2020-09 | .................... | .................... | .................... | .................... | .................... | .................... |
| V | 4633 | hCoV-19/Japan/PG-6940/2020\|EPI_ISL_688045\|2020-09 | .................... | .................... | .................... | .................... | .................... | .................... |
| V | 4634 | hCoV-19/Japan/PG-6941/2020\|EPI_ISL_688046\|2020-09 | .................... | .................... | .................... | .................... | .................... | .................... |
| V | 4635 | hCoV-19/Japan/PG-6942/2020\|EPI_ISL_688047\|2020-09 | .................... | .................... | .................... | .................... | .................... | .................... |
| V | 4636 | hCoV-19/Japan/PG-6943/2020\|EPI_ISL_688048\|2020 | .................... | .................... | .................... | .................... | .................... | .................... |
| V | 4637 | hCoV-19/Japan/PG-6944/2020\|EPI_ISL_688049\|2020 | .................... | .................... | .................... | .................... | .................... | .................... |

| | | | | | | | | |
|---|---|---|---|---|---|---|---|---|
| V | 4638 | hCoV-19/Japan/PG-6945/2020\|EPI_ISL_688050\|2020 | .................... | .................... | ..................... | .................... | .................... | .................... |
| V | 4639 | hCoV-19/Japan/PG-6946/2020\|EPI_ISL_688051\|2020 | .................... | .................... | ..................... | .................... | .................... | .................... |
| V | 4640 | hCoV-19/Japan/PG-6947/2020\|EPI_ISL_688052\|2020 | .................... | .................... | ..................... | .................... | .................... | .................... |
| V | 4641 | hCoV-19/Japan/PG-6948/2020\|EPI_ISL_688053\|2020 | .................... | .................... | ..................... | .................... | .................... | .................... |
| V | 4642 | hCoV-19/Japan/PG-6949/2020\|EPI_ISL_688054\|2020 | .................... | .................... | ..................... | .................... | .................... | .................... |
| V | 4643 | hCoV-19/Japan/PG-6950/2020\|EPI_ISL_688055\|2020 | .................... | .................... | ..................... | .................... | .................... | .................... |
| V | 4644 | hCoV-19/Japan/PG-6951/2020\|EPI_ISL_688056\|2020-07 | .................... | .................... | ..................... | .................... | .................... | .................... |
| V | 4645 | hCoV-19/Japan/PG-6952/2020\|EPI_ISL_688057\|2020-07 | .................... | .................... | ..................... | .................... | .................... | .................... |
| V | 4646 | hCoV-19/Japan/PG-6953/2020\|EPI_ISL_688058\|2020-07 | .................... | .................... | ..................... | .................... | .................... | .................... |
| V | 4647 | hCoV-19/Japan/PG-6954/2020\|EPI_ISL_688059\|2020-07 | .................... | .................... | ..................... | .................... | .................... | .................... |
| V | 4648 | hCoV-19/Japan/PG-6955/2020\|EPI_ISL_688060\|2020-07 | .................... | .................... | ..................... | .................... | .................... | .................... |
| V | 4649 | hCoV-19/Japan/PG-6956/2020\|EPI_ISL_688061\|2020-07 | .................... | .................... | ..................... | .................... | .................... | .................... |
| V | 4650 | hCoV-19/Japan/PG-6957/2020\|EPI_ISL_688062\|2020-07 | .................... | .................... | ..................... | .................... | .................... | .................... |
| V | 4651 | hCoV-19/Japan/PG-6958/2020\|EPI_ISL_688063\|2020-07 | .................... | .................... | ..................... | .................... | .................... | .................... |
| V | 4652 | hCoV-19/Japan/PG-6959/2020\|EPI_ISL_688064\|2020-07 | .................... | .................... | ..................... | .................... | .................... | .................... |
| V | 4653 | hCoV-19/Japan/PG-6960/2020\|EPI_ISL_688065\|2020-07 | .................... | .................... | ..................... | .................... | .................... | .................... |
| V | 4654 | hCoV-19/Japan/PG-6961/2020\|EPI_ISL_688066\|2020-07 | .................... | .................... | ..................... | .................... | .................... | .................... |
| V | 4655 | hCoV-19/Japan/PG-6962/2020\|EPI_ISL_688067\|2020-07 | .................... | .................... | ..................... | .................... | .................... | .................... |
| V | 4656 | hCoV-19/Japan/PG-6963/2020\|EPI_ISL_688068\|2020-07 | .................... | .................... | ..................... | .................... | .................... | .................... |
| V | 4657 | hCoV-19/Japan/PG-6964/2020\|EPI_ISL_688069\|2020-07 | .................... | .................... | ..................... | .................... | .................... | .................... |
| V | 4658 | hCoV-19/Japan/PG-6965/2020\|EPI_ISL_688070\|2020-07 | .................... | .................... | ..................... | .................... | .................... | .................... |
| V | 4659 | hCoV-19/Japan/PG-6966/2020\|EPI_ISL_688071\|2020-07 | .................... | .................... | ..................... | .................... | .................... | .................... |
| V | 4660 | hCoV-19/Japan/PG-6967/2020\|EPI_ISL_688072\|2020-07 | .................... | .................... | ..................... | .................... | .................... | .................... |
| V | 4661 | hCoV-19/Japan/PG-6968/2020\|EPI_ISL_688073\|2020-07 | .................... | .................... | ..................... | .................... | .................... | .................... |
| V | 4662 | hCoV-19/Japan/PG-6969/2020\|EPI_ISL_688074\|2020-07 | .................... | .................... | ..................... | .................... | .................... | .................... |
| V | 4663 | hCoV-19/Japan/PG-6970/2020\|EPI_ISL_688075\|2020-07 | .................... | .................... | ..................... | .................... | .................... | .................... |
| V | 4664 | hCoV-19/Japan/PG-6971/2020\|EPI_ISL_688076\|2020-07 | .................... | .................... | ..................... | .................... | .................... | .................... |
| V | 4665 | hCoV-19/Japan/PG-6972/2020\|EPI_ISL_688077\|2020-07 | .................... | .................... | ..................... | .................... | .................... | .................... |
| V | 4666 | hCoV-19/Japan/PG-6973/2020\|EPI_ISL_688078\|2020-07 | .................... | .................... | ..................... | .................... | .................... | .................... |
| V | 4667 | hCoV-19/Japan/PG-6974/2020\|EPI_ISL_688079\|2020-07 | .................... | .................... | ..................... | .................... | .................... | .................... |
| V | 4668 | hCoV-19/Japan/PG-6188/2020\|EPI_ISL_689300\|2020-08 | .................... | .................... | ..................... | .................... | .................... | .................... |
| V | 4669 | hCoV-19/Japan/PG-6189/2020\|EPI_ISL_689301\|2020-08 | .................... | .................... | ..................... | .................... | .................... | .................... |
| V | 4670 | hCoV-19/Japan/PG-6190/2020\|EPI_ISL_689302\|2020-08 | .................... | .................... | ..................... | .................... | .................... | .................... |
| V | 4671 | hCoV-19/Japan/PG-6191/2020\|EPI_ISL_689303\|2020-08 | .................... | .................... | ..................... | .................... | .................... | .................... |
| V | 4672 | hCoV-19/Japan/PG-6239/2020\|EPI_ISL_689351\|2020-08 | .................... | .................... | ..................... | .................... | .................... | .................... |
| V | 4673 | hCoV-19/Japan/PG-6240/2020\|EPI_ISL_689352\|2020-08 | .................... | .................... | ..................... | .................... | .................... | .................... |
| V | 4674 | hCoV-19/Japan/PG-6241/2020\|EPI_ISL_689353\|2020-08 | .................... | .................... | ..................... | .................... | .................... | .................... |
| V | 4675 | hCoV-19/Japan/PG-6243/2020\|EPI_ISL_689354\|2020-08 | .................... | .................... | ..................... | .................... | .................... | .................... |
| V | 4676 | hCoV-19/Japan/PG-6244/2020\|EPI_ISL_689355\|2020-08 | .................... | .................... | ..................... | .................... | .................... | .................... |
| V | 4677 | hCoV-19/Japan/PG-6245/2020\|EPI_ISL_689356\|2020-08 | .................... | .................... | ..................... | .................... | .................... | .................... |
| V | 4678 | hCoV-19/Japan/PG-6246/2020\|EPI_ISL_689357\|2020-08 | .................... | .................... | ..................... | .................... | .................... | .................... |
| V | 4679 | hCoV-19/Japan/PG-6247/2020\|EPI_ISL_689358\|2020-08 | .................... | .................... | ..................... | .................... | .................... | .................... |
| V | 4680 | hCoV-19/Japan/PG-6248/2020\|EPI_ISL_689359\|2020-08 | .................... | .................... | ..................... | .................... | .................... | .................... |
| V | 4681 | hCoV-19/Japan/PG-6249/2020\|EPI_ISL_689360\|2020-08 | .................... | .................... | ..................... | .................... | .................... | .................... |
| V | 4682 | hCoV-19/Japan/PG-6250/2020\|EPI_ISL_689361\|2020-08 | .................... | .................... | ..................... | .................... | .................... | .................... |
| V | 4683 | hCoV-19/Japan/PG-6251/2020\|EPI_ISL_689362\|2020-08 | .................... | .................... | ..................... | .................... | .................... | .................... |
| V | 4684 | hCoV-19/Japan/PG-6252/2020\|EPI_ISL_689363\|2020-08 | .................... | .................... | ..................... | .................... | .................... | .................... |
| V | 4685 | hCoV-19/Japan/PG-6253/2020\|EPI_ISL_689364\|2020-08 | .................... | .................... | ..................... | .................... | .................... | .................... |
| V | 4686 | hCoV-19/Japan/PG-6254/2020\|EPI_ISL_689365\|2020-08 | .................... | .................... | ..................... | .................... | .................... | .................... |
| V | 4687 | hCoV-19/Japan/PG-6255/2020\|EPI_ISL_689366\|2020-08 | .................... | .................... | ..................... | .................... | .................... | .................... |
| V | 4688 | hCoV-19/Japan/PG-6256/2020\|EPI_ISL_689367\|2020-08 | .................... | .................... | ..................... | .................... | .................... | .................... |
| V | 4689 | hCoV-19/Japan/PG-6257/2020\|EPI_ISL_689368\|2020-08 | .................... | .................... | ..................... | .................... | .................... | .................... |
| V | 4690 | hCoV-19/Japan/PG-6258/2020\|EPI_ISL_689369\|2020-08 | .................... | .................... | ..................... | .................... | .................... | .................... |
| V | 4691 | hCoV-19/Japan/PG-6259/2020\|EPI_ISL_689370\|2020-08 | .................... | .................... | ..................... | .................... | .................... | .................... |
| V | 4692 | hCoV-19/Japan/PG-6260/2020\|EPI_ISL_689371\|2020-08 | .................... | .................... | ..................... | .................... | .................... | .................... |
| V | 4693 | hCoV-19/Japan/PG-6261/2020\|EPI_ISL_689372\|2020-08 | .................... | .................... | ..................... | .................... | .................... | .................... |
| V | 4694 | hCoV-19/Japan/PG-6262/2020\|EPI_ISL_689373\|2020-08 | .................... | .................... | ..................... | .................... | .................... | .................... |
| V | 4695 | hCoV-19/Japan/PG-6263/2020\|EPI_ISL_689374\|2020-08 | .................... | .................... | ..................... | .................... | .................... | .................... |
| V | 4696 | hCoV-19/Japan/PG-6264/2020\|EPI_ISL_689375\|2020-08 | .................... | .................... | ..................... | .................... | .................... | .................... |
| V | 4697 | hCoV-19/Japan/PG-6265/2020\|EPI_ISL_689376\|2020-08 | .................... | .................... | ..................... | .................... | .................... | .................... |
| V | 4698 | hCoV-19/Japan/PG-6266/2020\|EPI_ISL_689377\|2020-08 | .................... | .................... | ..................... | .................... | .................... | .................... |
| V | 4699 | hCoV-19/Japan/PG-6267/2020\|EPI_ISL_689378\|2020-08 | .................... | .................... | ..................... | .................... | .................... | .................... |
| V | 4700 | hCoV-19/Japan/PG-6268/2020\|EPI_ISL_689379\|2020-08 | .................... | .................... | ..................... | .................... | .................... | .................... |
| V | 4701 | hCoV-19/Japan/PG-6269/2020\|EPI_ISL_689380\|2020-08 | .................... | .................... | ..................... | .................... | .................... | .................... |
| V | 4702 | hCoV-19/Japan/PG-6270/2020\|EPI_ISL_689381\|2020-08 | .................... | .................... | ..................... | .................... | .................... | .................... |
| V | 4703 | hCoV-19/Japan/PG-6271/2020\|EPI_ISL_689382\|2020-08 | .................... | .................... | ..................... | .................... | .................... | .................... |
| V | 4704 | hCoV-19/Japan/PG-6272/2020\|EPI_ISL_689383\|2020-08 | .................... | .................... | ..................... | .................... | .................... | .................... |
| V | 4705 | hCoV-19/Japan/PG-6273/2020\|EPI_ISL_689384\|2020-08 | .................... | .................... | ..................... | .................... | .................... | .................... |
| V | 4706 | hCoV-19/Japan/PG-6274/2020\|EPI_ISL_689385\|2020-08 | .................... | .................... | ..................... | .................... | .................... | .................... |
| V | 4707 | hCoV-19/Japan/PG-6275/2020\|EPI_ISL_689386\|2020-08 | .................... | .................... | ..................... | .................... | .................... | .................... |
| V | 4708 | hCoV-19/Japan/PG-6276/2020\|EPI_ISL_689387\|2020-08 | .................... | .................... | ..................... | .................... | .................... | .................... |
| V | 4709 | hCoV-19/Japan/PG-6277/2020\|EPI_ISL_689388\|2020-08 | .................... | .................... | ..................... | .................... | .................... | .................... |
| V | 4710 | hCoV-19/Japan/PG-6278/2020\|EPI_ISL_689389\|2020-08 | .................... | .................... | ..................... | .................... | .................... | .................... |
| V | 4711 | hCoV-19/Japan/PG-6279/2020\|EPI_ISL_689390\|2020-08 | .................... | .................... | ..................... | .................... | .................... | .................... |
| V | 4712 | hCoV-19/Japan/PG-6280/2020\|EPI_ISL_689391\|2020-08 | .................... | .................... | ..................... | .................... | .................... | .................... |
| V | 4713 | hCoV-19/Japan/PG-6281/2020\|EPI_ISL_689392\|2020-08 | .................... | .................... | ..................... | .................... | .................... | .................... |
| V | 4714 | hCoV-19/Japan/PG-6282/2020\|EPI_ISL_689393\|2020-08 | .................... | .................... | ..................... | .................... | .................... | .................... |
| V | 4715 | hCoV-19/Japan/PG-6283/2020\|EPI_ISL_689394\|2020-08 | .................... | .................... | ..................... | .................... | .................... | .................... |
| V | 4716 | hCoV-19/Japan/PG-6284/2020\|EPI_ISL_689395\|2020-07 | .................... | .................... | ..................... | .................... | .................... | .................... |
| V | 4717 | hCoV-19/Japan/PG-6285/2020\|EPI_ISL_689396\|2020-07 | .................... | .................... | ..................... | .................... | .................... | .................... |

V 4718 hCoV-19/Japan/PG-6286/2020|EPI_ISL_689397|2020-07 ···················· ···················· ···················· ···················· ···················· ····················
V 4719 hCoV-19/Japan/PG-6287/2020|EPI_ISL_689398|2020-07 ···················· ···················· ···················· ···················· ···················· ····················
V 4720 hCoV-19/Japan/PG-6288/2020|EPI_ISL_689399|2020-07 ···················· ···················· ···················· ···················· ···················· ····················
V 4721 hCoV-19/Japan/PG-5227/2020|EPI_ISL_690330|2020-08 ···················· ···················· ···················· ···················· ···················· ····················
V 4722 hCoV-19/Japan/PG-5228/2020|EPI_ISL_690331|2020-08 ···················· ···················· ···················· ···················· ···················· ····················
V 4723 hCoV-19/Japan/PG-5229/2020|EPI_ISL_690332|2020-08 ···················· ···················· ···················· ···················· ···················· ····················
V 4724 hCoV-19/Japan/PG-5230/2020|EPI_ISL_690333|2020-08 ···················· ···················· ···················· ···················· ···················· ····················
V 4725 hCoV-19/Japan/PG-5231/2020|EPI_ISL_690334|2020-08 ···················· ···················· ···················· ···················· ···················· ····················
V 4726 hCoV-19/Japan/PG-5232/2020|EPI_ISL_690335|2020-08 ···················· ···················· ···················· ···················· ···················· ····················
V 4727 hCoV-19/Japan/PG-5233/2020|EPI_ISL_690336|2020-08 ···················· ···················· ···················· ···················· ···················· ····················
V 4728 hCoV-19/Japan/PG-5236/2020|EPI_ISL_690339|2020-08 ···················· ···················· ···················· ···················· ···················· ····················
V 4729 hCoV-19/Japan/PG-5237/2020|EPI_ISL_690340|2020-08 ···················· ···················· ···················· ···················· ···················· ····················
V 4730 hCoV-19/Japan/PG-5238/2020|EPI_ISL_690341|2020-08 ···················· ···················· ···················· ···················· ···················· ····················
V 4731 hCoV-19/Japan/PG-5239/2020|EPI_ISL_690342|2020-08 ···················· ···················· ···················· ···················· ···················· ····················
V 4732 hCoV-19/Japan/PG-5240/2020|EPI_ISL_690343|2020-08 ···················· ···················· ···················· ···················· ···················· ····················
V 4733 hCoV-19/Japan/PG-5241/2020|EPI_ISL_690344|2020-08 ···················· ···················· ···················· ···················· ···················· ····················
V 4734 hCoV-19/Japan/PG-5242/2020|EPI_ISL_690345|2020-08 ···················· ···················· ···················· ···················· ···················· ····················
V 4735 hCoV-19/Japan/PG-5243/2020|EPI_ISL_690346|2020-08 ···················· ···················· ···················· ···················· ···················· ····················
V 4736 hCoV-19/Japan/PG-5244/2020|EPI_ISL_690347|2020-08 ···················· ···················· ···················· ···················· ···················· ····················
V 4737 hCoV-19/Japan/PG-5245/2020|EPI_ISL_690348|2020-08 ···················· ···················· ···················· ···················· ···················· ····················
V 4738 hCoV-19/Japan/PG-5246/2020|EPI_ISL_690349|2020-07 ···················· ···················· ···················· ···················· ···················· ····················
V 4739 hCoV-19/Japan/PG-5247/2020|EPI_ISL_690350|2020-07 ···················· ···················· ···················· ···················· ···················· ····················
V 4740 hCoV-19/Japan/PG-5248/2020|EPI_ISL_690351|2020-07 ···················· ···················· ···················· ···················· ···················· ····················
V 4741 hCoV-19/Japan/PG-5249/2020|EPI_ISL_690352|2020-07 ···················· ···················· ···················· ···················· ···················· ····················
V 4742 hCoV-19/Japan/PG-5250/2020|EPI_ISL_690353|2020-07 ···················· ···················· ···················· ···················· ···················· ····················
V 4743 hCoV-19/Japan/PG-5251/2020|EPI_ISL_690354|2020-07 ···················· ···················· ···················· ···················· ···················· ····················
V 4744 hCoV-19/Japan/PG-5252/2020|EPI_ISL_690355|2020-07 ···················· ···················· ···················· ···················· ···················· ····················
V 4745 hCoV-19/Japan/PG-5253/2020|EPI_ISL_690356|2020-08 ···················· ···················· ···················· ···················· ···················· ····················
V 4746 hCoV-19/Japan/PG-5254/2020|EPI_ISL_690357|2020-07 ···················· ···················· ···················· ···················· ···················· ····················
V 4747 hCoV-19/Japan/PG-5255/2020|EPI_ISL_690358|2020-07 ···················· ···················· ···················· ···················· ···················· ····················
V 4748 hCoV-19/Japan/PG-5256/2020|EPI_ISL_690359|2020-07 ···················· ···················· ···················· ···················· ···················· ····················
V 4749 hCoV-19/Japan/PG-5257/2020|EPI_ISL_690360|2020-07 ···················· ···················· ···················· ···················· ···················· ····················
V 4750 hCoV-19/Japan/PG-5258/2020|EPI_ISL_690361|2020-07 ···················· ···················· ···················· ···················· ···················· ····················
V 4751 hCoV-19/Japan/PG-5259/2020|EPI_ISL_690362|2020-08 ···················· ···················· ···················· ···················· ···················· ····················
V 4752 hCoV-19/Japan/PG-5260/2020|EPI_ISL_690363|2020-08 ···················· ···················· ···················· ···················· ···················· ····················
V 4753 hCoV-19/Japan/PG-5261/2020|EPI_ISL_690364|2020-08 ···················· ···················· ···················· ···················· ···················· ····················
V 4754 hCoV-19/Japan/PG-5262/2020|EPI_ISL_690365|2020-08 ···················· ···················· ···················· ···················· ···················· ····················
V 4755 hCoV-19/Japan/PG-5263/2020|EPI_ISL_690366|2020-08 ···················· ···················· ···················· ···················· ···················· ····················
V 4756 hCoV-19/Japan/PG-5264/2020|EPI_ISL_690367|2020-07 ···················· ···················· ···················· ···················· ···················· ····················
V 4757 hCoV-19/Japan/PG-5265/2020|EPI_ISL_690368|2020-07 ···················· ···················· ···················· ···················· ···················· ····················
V 4758 hCoV-19/Japan/PG-5266/2020|EPI_ISL_690369|2020-08 ···················· ···················· ···················· ···················· ···················· ····················
V 4759 hCoV-19/Japan/PG-5267/2020|EPI_ISL_690370|2020-08 ···················· ···················· ···················· ···················· ···················· ····················
V 4760 hCoV-19/Japan/PG-5268/2020|EPI_ISL_690371|2020-08 ···················· ···················· ···················· ···················· ···················· ····················
V 4761 hCoV-19/Japan/PG-5269/2020|EPI_ISL_690372|2020-08 ···················· ···················· ···················· ···················· ···················· ····················
V 4762 hCoV-19/Japan/PG-5270/2020|EPI_ISL_690373|2020-08 ···················· ···················· ···················· ···················· ···················· ····················
V 4763 hCoV-19/Japan/PG-5271/2020|EPI_ISL_690374|2020-08 ···················· ···················· ···················· ···················· ···················· ····················
V 4764 hCoV-19/Japan/PG-5272/2020|EPI_ISL_690375|2020-08 ···················· ···················· ···················· ···················· ···················· ····················
V 4765 hCoV-19/Japan/PG-5273/2020|EPI_ISL_690376|2020-08 ···················· ···················· ···················· ···················· ···················· ····················
V 4766 hCoV-19/Japan/PG-5274/2020|EPI_ISL_690377|2020-08 ···················· ···················· ···················· ···················· ···················· ····················
V 4767 hCoV-19/Japan/PG-5275/2020|EPI_ISL_690378|2020-08 ···················· ···················· ···················· ···················· ···················· ····················
V 4768 hCoV-19/Japan/PG-5276/2020|EPI_ISL_690379|2020-08 ···················· ···················· ···················· ···················· ···················· ····················
V 4769 hCoV-19/Japan/PG-5277/2020|EPI_ISL_690380|2020-08 ···················· ···················· ···················· ···················· ···················· ····················
V 4770 hCoV-19/Japan/PG-5278/2020|EPI_ISL_690381|2020-08 ···················· ···················· ···················· ···················· ···················· ····················
V 4771 hCoV-19/Japan/PG-5279/2020|EPI_ISL_690382|2020-08 ···················· ···················· ···················· ···················· ···················· ····················
V 4772 hCoV-19/Japan/PG-5280/2020|EPI_ISL_690383|2020-08 ···················· ···················· ···················· ···················· ···················· ····················
V 4773 hCoV-19/Japan/PG-5281/2020|EPI_ISL_690384|2020-08 ···················· ···················· ···················· ···················· ···················· ····················
V 4774 hCoV-19/Japan/PG-5282/2020|EPI_ISL_690385|2020-07 ···················· ···················· ···················· ···················· ···················· ····················
V 4775 hCoV-19/Japan/PG-5283/2020|EPI_ISL_690386|2020-07 ···················· ···················· ···················· ···················· ···················· ····················
V 4777 hCoV-19/Japan/PG-5285/2020|EPI_ISL_690388|2020-07 ···················· ···················· ···················· ···················· ···················· ····················
V 4778 hCoV-19/Japan/PG-5286/2020|EPI_ISL_690389|2020-07 ···················· ···················· ···················· ···················· ···················· ····················
V 4779 hCoV-19/Japan/PG-5287/2020|EPI_ISL_690390|2020-08 ···················· ···················· ···················· ···················· ···················· ····················
V 4780 hCoV-19/Japan/PG-5288/2020|EPI_ISL_690391|2020-08 ···················· ···················· ···················· ···················· ···················· ····················
V 4781 hCoV-19/Japan/PG-5289/2020|EPI_ISL_690392|2020-08 ···················· ···················· ···················· ···················· ···················· ····················
V 4782 hCoV-19/Japan/PG-5290/2020|EPI_ISL_690393|2020-08 ···················· ···················· ···················· ···················· ···················· ····················
V 4783 hCoV-19/Japan/PG-5291/2020|EPI_ISL_690394|2020-08 ···················· ···················· ···················· ···················· ···················· ····················
V 4784 hCoV-19/Japan/PG-3491/2020|EPI_ISL_691600|2020-07 ···················· ···················· ···················· ···················· ···················· ····················
V 4785 hCoV-19/Japan/PG-3492/2020|EPI_ISL_691601|2020-07 ···················· ···················· ···················· ···················· ···················· ····················
V 4786 hCoV-19/Japan/PG-3493/2020|EPI_ISL_691602|2020-07 ···················· ···················· ···················· ···················· ···················· ····················
V 4787 hCoV-19/Japan/PG-3494/2020|EPI_ISL_691603|2020-07 ···················· ···················· ···················· ···················· ···················· ····················
V 4788 hCoV-19/Japan/PG-3495/2020|EPI_ISL_691604|2020-07 ···················· ···················· ···················· ···················· ···················· ····················
V 4789 hCoV-19/Japan/PG-3496/2020|EPI_ISL_691605|2020-07 ···················· ···················· ···················· ···················· ···················· ····················
V 4790 hCoV-19/Japan/PG-3497/2020|EPI_ISL_691606|2020-07 ···················· ···················· ···················· ···················· ···················· ····················
V 4791 hCoV-19/Japan/PG-3498/2020|EPI_ISL_691607|2020-07 ···················· ···················· ···················· ···················· ···················· ····················
V 4792 hCoV-19/Japan/PG-3499/2020|EPI_ISL_691608|2020-07 ···················· ···················· ···················· ···················· ···················· ····················
V 4793 hCoV-19/Japan/PG-3500/2020|EPI_ISL_691609|2020-07 ···················· ···················· ···················· ···················· ···················· ····················
V 4795 hCoV-19/Spain/IB-IBV-99010760/2020|EPI_ISL_691662| ···················· ···················· ···················· ···················· ···················· ····················
V 4796 hCoV-19/Spain/IB-IBV-99010763/2020|EPI_ISL_691663| ···················· ···················· ···················· ···················· ···················· ····················
V 4797 hCoV-19/Spain/IB-IBV-99010764/2020|EPI_ISL_691664| ···················· ···················· ···················· ···················· ···················· ····················
V 4798 hCoV-19/Spain/IB-IBV-99010768/2020|EPI_ISL_691665| ···················· ···················· ···················· ···················· ·····T·············· ····················
V 4799 hCoV-19/Spain/IB-IBV-99010769/2020|EPI_ISL_691666| ···················· ···················· ···················· ···················· ···················· ····················

| | | | | | | | |
|---|---|---|---|---|---|---|---|
| V | 4800 hCoV-19/Spain/IB-IBV-99010774/2020\|EPI_ISL_691667\| | ···················· | ···················· | ···················· | ···················· | ···················· | ···················· |
| V | 4801 hCoV-19/Spain/IB-IBV-99010775/2020\|EPI_ISL_691668\| | ···················· | ···················· | ···················· | ···················· | ···················· | ···················· |
| V | 4802 hCoV-19/Spain/IB-IBV-99010777/2020\|EPI_ISL_691669\| | ···················· | ···················· | ···················· | ···················· | ···················· | ···················· |
| V | 4803 hCoV-19/Spain/IB-IBV-99010778/2020\|EPI_ISL_691670\| | ···················· | ···················· | ···················· | ···················· | ···················· | ···················· |
| V | 4804 hCoV-19/Spain/IB-IBV-99010780/2020\|EPI_ISL_691671\| | ···················· | ···················· | ···················· | ···················· | ···················· | ···················· |
| V | 4805 hCoV-19/Spain/IB-IBV-99010782/2020\|EPI_ISL_691672\| | ···················· | ···················· | ···················· | ···················· | ···················· | ···················· |
| V | 4806 hCoV-19/Spain/IB-IBV-99010783/2020\|EPI_ISL_691673\| | ···················· | ···················· | ···················· | ···················· | ···················· | ···················· |
| V | 4807 hCoV-19/Spain/IB-IBV-99010785/2020\|EPI_ISL_691674\| | ···················· | ···················· | ···················· | ···················· | ······T············· | ···················· |
| V | 4808 hCoV-19/Spain/IB-IBV-99010786/2020\|EPI_ISL_691675\| | ···················· | ···················· | ···················· | ···················· | ···················· | ···················· |
| V | 4809 hCoV-19/Spain/IB-IBV-99010788/2020\|EPI_ISL_691676\| | ···················· | ···················· | ···················· | ···················· | ···················· | ···················· |
| V | 4810 hCoV-19/Spain/IB-IBV-99010790/2020\|EPI_ISL_691677\| | ···················· | ···················· | ···················· | ···················· | ···················· | ···················· |
| V | 4813 hCoV-19/Spain/IB-IBV-99010811/2020\|EPI_ISL_691680\| | ···················· | ···················· | ···················· | ···················· | ·NN················· | ···················· |
| V | 4814 hCoV-19/Spain/CM-ISCIII-201368/2020\|EPI_ISL_691681 | ···················· | ···················· | ···················· | ···················· | ···················· | ···················· |
| V | 4815 hCoV-19/Spain/CM-ISCIII-2010312/2020\|EPI_ISL_69168 | ···················· | ···················· | ···················· | ···················· | ····T··············· | ···················· |
| V | 4816 hCoV-19/Spain/CM-ISCIII-2010317/2020\|EPI_ISL_69168 | ···················· | ···················· | ···················· | ···················· | ···················· | ···················· |
| V | 4817 hCoV-19/Spain/CM-ISCIII-2010740/2020\|EPI_ISL_69168 | ···················· | ···················· | ···················· | ···················· | ···················· | ···················· |
| V | 4818 hCoV-19/Spain/CL-ISCIII-2016156/2020\|EPI_ISL_69168 | ···················· | ···················· | ···················· | ···················· | ···················· | ···················· |
| V | 4819 hCoV-19/Spain/CE-ISCIII-2016496/2020\|EPI_ISL_69168 | ···················· | ···················· | ···················· | ···················· | ···················· | ···················· |
| V | 4820 hCoV-19/Spain/CE-ISCIII-2016502/2020\|EPI_ISL_69168 | ···················· | ·A·················· | ···················· | ···················· | ···················· | ···················· |
| V | 4821 hCoV-19/Spain/CE-ISCIII-2016503/2020\|EPI_ISL_69168 | ···················· | ···················· | ···················· | ···················· | ···················· | ···················· |
| V | 4822 hCoV-19/Spain/CE-ISCIII-2016504/2020\|EPI_ISL_69168 | ···················· | ···················· | ···················· | ···················· | ···················· | ···················· |
| V | 4823 hCoV-19/Spain/CE-ISCIII-2016506/2020\|EPI_ISL_69169 | ···················· | ···················· | ···················· | ···················· | ···················· | ···················· |
| V | 4824 hCoV-19/Spain/CE-ISCIII-2016510/2020\|EPI_ISL_69169 | ···················· | ···················· | ···················· | ···················· | ···················· | ···················· |
| V | 4825 hCoV-19/Spain/CE-ISCIII-2016511/2020\|EPI_ISL_69169 | ···················· | ···················· | ···················· | ···················· | ···················· | ···················· |
| V | 4826 hCoV-19/Spain/CE-ISCIII-2016513/2020\|EPI_ISL_69169 | ···················· | ···················· | ···················· | ···················· | ···················· | ···················· |
| V | 4827 hCoV-19/Spain/CE-ISCIII-2016514/2020\|EPI_ISL_69169 | ···················· | ···················· | ···················· | ···················· | ···················· | ···················· |
| V | 4828 hCoV-19/Spain/CE-ISCIII-2016516/2020\|EPI_ISL_69169 | ···················· | ···················· | ···················· | ···················· | ···················· | ···················· |
| V | 4829 hCoV-19/Spain/CE-ISCIII-2016517/2020\|EPI_ISL_69169 | ···················· | ···················· | ···················· | ···················· | ···················· | ···················· |
| V | 4830 hCoV-19/Spain/CE-ISCIII-2016518/2020\|EPI_ISL_69169 | ···················· | ···················· | ···················· | ···················· | ···················· | ···················· |
| V | 4831 hCoV-19/Spain/CE-ISCIII-2016519/2020\|EPI_ISL_69169 | ···················· | ···················· | ···················· | ···················· | ···················· | ···················· |
| V | 4832 hCoV-19/Spain/CE-ISCIII-2016520/2020\|EPI_ISL_69169 | ···················· | ···················· | ···················· | ···················· | ···················· | ···················· |
| V | 4833 hCoV-19/USA/MA-MASPHL-00800/2020\|EPI_ISL_692900\|20 | ···················· | ···················· | ···················· | ···················· | ···················· | ···················· |
| V | 4834 hCoV-19/USA/MA-MASPHL-00798/2020\|EPI_ISL_692901\|20 | ···················· | ···················· | ···················· | ···················· | ···················· | ···················· |
| V | 4835 hCoV-19/USA/MA-MASPHL-00795/2020\|EPI_ISL_692903\|20 | ···················· | ···················· | ···················· | ···················· | ···················· | ···················· |
| V | 4836 hCoV-19/USA/MA-MASPHL-00789/2020\|EPI_ISL_692909\|20 | ···················· | ·············A······ | ···················· | ···················· | ···················· | ···················· |
| V | 4837 hCoV-19/USA/MA-MASPHL-00788/2020\|EPI_ISL_692910\|20 | ···················· | ···················· | ···················· | ···················· | ···················· | ···················· |
| V | 4838 hCoV-19/USA/MA-MASPHL-00786/2020\|EPI_ISL_692911\|20 | ···················· | ···················· | ···················· | ···················· | ···················· | ···················· |
| V | 4839 hCoV-19/USA/MA-MASPHL-00785/2020\|EPI_ISL_692912\|20 | ···················· | ·A·················· | ···················· | ···················· | ···················· | ···················· |
| V | 4840 hCoV-19/USA/MA-MASPHL-00784/2020\|EPI_ISL_692913\|20 | ···················· | ···················· | ···················· | ···················· | ···················· | ···················· |
| V | 4841 hCoV-19/USA/MA-MASPHL-00781/2020\|EPI_ISL_692915\|20 | ···················· | ···················· | ···················· | ···················· | ···················· | ···················· |
| V | 4842 hCoV-19/USA/MA-MASPHL-00777/2020\|EPI_ISL_692918\|20 | ···················· | ···················· | ···················· | ···················· | ···················· | ···················· |
| V | 4843 hCoV-19/USA/MA-MASPHL-00776/2020\|EPI_ISL_692919\|20 | ···················· | ···················· | ···················· | ···················· | ···················· | ···················· |
| V | 4844 hCoV-19/USA/MA-MASPHL-00774/2020\|EPI_ISL_692921\|20 | ···················· | ···················· | ···················· | ···················· | ···················· | ···················· |
| V | 4845 hCoV-19/USA/MA-MASPHL-00773/2020\|EPI_ISL_692922\|20 | ···················· | ···················· | ···················· | ···················· | ···················· | ···················· |
| V | 4846 hCoV-19/USA/MA-MASPHL-00772/2020\|EPI_ISL_692923\|20 | ···················· | ···················· | ···················· | ···················· | ···················· | ···················· |
| V | 4847 hCoV-19/USA/MA-MASPHL-00771/2020\|EPI_ISL_692924\|20 | ···················· | ···················· | ···················· | ···················· | ···················· | ···················· |
| V | 4848 hCoV-19/USA/MA-MASPHL-00748/2020\|EPI_ISL_692926\|20 | ···················· | ···················· | ···················· | ···················· | ···················· | ···················· |
| V | 4849 hCoV-19/USA/MA-MASPHL-00746/2020\|EPI_ISL_692927\|20 | ···················· | ···················· | ···················· | ···················· | ···················· | ···················· |
| V | 4850 hCoV-19/USA/MA-MASPHL-00745/2020\|EPI_ISL_692928\|20 | ···················· | ···················· | ···················· | ···················· | ···················· | ···················· |
| V | 4851 hCoV-19/USA/MA-MASPHL-00744/2020\|EPI_ISL_692929\|20 | ···················· | ···················· | ···················· | ···················· | ···················· | ···················· |
| V | 4852 hCoV-19/USA/MA-MASPHL-00743/2020\|EPI_ISL_692930\|20 | ···················· | ···················· | ···················· | ···················· | ···················· | ···················· |
| V | 4853 hCoV-19/USA/MA-MASPHL-00742/2020\|EPI_ISL_692931\|20 | ···················· | ···················· | ···················· | ···················· | ···················· | ···················· |
| V | 4854 hCoV-19/USA/MA-MASPHL-00739/2020\|EPI_ISL_692933\|20 | ···················· | ···················· | ···················· | ···················· | ···················· | ···················· |
| V | 4855 hCoV-19/USA/MA-MASPHL-00737/2020\|EPI_ISL_692935\|20 | ···················· | ···················· | ···················· | ···················· | ···················· | ···················· |
| V | 4856 hCoV-19/USA/MA-MASPHL-00735/2020\|EPI_ISL_692937\|20 | ···················· | ···················· | ···················· | ···················· | ···················· | ···················· |
| V | 4857 hCoV-19/USA/MA-MASPHL-00733/2020\|EPI_ISL_692939\|20 | ···················· | ···················· | ···················· | ···················· | ···················· | ···················· |
| V | 4858 hCoV-19/USA/MA-MASPHL-00728/2020\|EPI_ISL_692944\|20 | ···················· | ···················· | ···················· | ···················· | ···················· | ···················· |
| V | 4859 hCoV-19/USA/MA-MASPHL-00727/2020\|EPI_ISL_692945\|20 | ···················· | ···················· | ···················· | ···················· | ···················· | ···················· |
| V | 4860 hCoV-19/USA/MA-MASPHL-00726/2020\|EPI_ISL_692946\|20 | ···················· | ···················· | ···················· | ···················· | ···················· | ···················· |
| V | 4861 hCoV-19/USA/MA-MASPHL-00716/2020\|EPI_ISL_692947\|20 | ···················· | ···················· | ···················· | ···················· | ···················· | ···················· |
| V | 4862 hCoV-19/USA/MA-MASPHL-00699/2020\|EPI_ISL_692949\|20 | ···················· | ···················· | ···················· | ···················· | ···················· | ···················· |
| V | 4863 hCoV-19/USA/MA-MASPHL-00637/2020\|EPI_ISL_692992\|20 | ···················· | ···················· | ···················· | ···················· | ···················· | ···················· |
| V | 4864 hCoV-19/USA/MA-MASPHL-00632/2020\|EPI_ISL_692996\|20 | ···················· | ···················· | ···················· | ···················· | ···················· | ···················· |
| V | 4865 hCoV-19/Japan/PG-8604/2020\|EPI_ISL_686200\|2020-08 | ···················· | ···················· | ···················· | ···················· | ···················· | ···················· |
| V | 4866 hCoV-19/Japan/PG-8605/2020\|EPI_ISL_686201\|2020-08 | ···················· | ···················· | ···················· | ···················· | ···················· | ···················· |
| V | 4867 hCoV-19/Japan/PG-8606/2020\|EPI_ISL_686202\|2020-08 | ···················· | ···················· | ···················· | ···················· | ···················· | ···················· |
| V | 4868 hCoV-19/Japan/PG-8607/2020\|EPI_ISL_686203\|2020-08 | ···················· | ···················· | ···················· | ···················· | ···················· | ···················· |
| V | 4869 hCoV-19/Japan/PG-8608/2020\|EPI_ISL_686204\|2020-08 | ···················· | ···················· | ···················· | ···················· | ···················· | ···················· |
| V | 4870 hCoV-19/Japan/PG-8609/2020\|EPI_ISL_686205\|2020-08 | ···················· | ···················· | ···················· | ···················· | ···················· | ···················· |
| V | 4871 hCoV-19/Japan/PG-8610/2020\|EPI_ISL_686206\|2020-08 | ···················· | ···················· | ···················· | ···················· | ···················· | ···················· |
| V | 4872 hCoV-19/Japan/PG-8611/2020\|EPI_ISL_686207\|2020-08 | ···················· | ···················· | ···················· | ···················· | ···················· | ···················· |
| V | 4873 hCoV-19/Japan/PG-8612/2020\|EPI_ISL_686208\|2020-08 | ···················· | ···················· | ···················· | ···················· | ···················· | ···················· |
| V | 4874 hCoV-19/Japan/PG-8613/2020\|EPI_ISL_686209\|2020-08 | ···················· | ···················· | ···················· | ···················· | ···················· | ···················· |
| V | 4875 hCoV-19/Japan/PG-8614/2020\|EPI_ISL_686210\|2020-08 | ···················· | ···················· | ···················· | ···················· | ···················· | ···················· |
| V | 4876 hCoV-19/Japan/PG-8615/2020\|EPI_ISL_686211\|2020-08 | ···················· | ···················· | ···················· | ···················· | ···················· | ···················· |
| V | 4877 hCoV-19/Japan/PG-8616/2020\|EPI_ISL_686212\|2020-08 | ···················· | ···················· | ···················· | ···················· | ···················· | ···················· |
| V | 4878 hCoV-19/Japan/PG-8617/2020\|EPI_ISL_686213\|2020-08 | ···················· | ···················· | ···················· | ···················· | ···················· | ···················· |
| V | 4879 hCoV-19/Japan/PG-8618/2020\|EPI_ISL_686214\|2020-09 | ···················· | ···················· | ···················· | ···················· | ···················· | ···················· |
| V | 4880 hCoV-19/Japan/PG-8411/2020\|EPI_ISL_687510\|2020-08 | ···················· | ···················· | ···················· | ···················· | ···················· | ···················· |
| V | 4881 hCoV-19/Japan/PG-8412/2020\|EPI_ISL_687511\|2020-08 | ···················· | ···················· | ···················· | ···················· | ···················· | ···················· |

| | | | | | | | | |
|---|---|---|---|---|---|---|---|---|
| V | 4882 | hCoV-19/Japan/PG-8413/2020\|EPI_ISL_687512\|2020-08 | .................... | .................... | ..................... | .................... | .................... | .................... |
| V | 4883 | hCoV-19/Japan/PG-8414/2020\|EPI_ISL_687513\|2020-08 | .................... | .................... | ..................... | .................... | .................... | .................... |
| V | 4884 | hCoV-19/Japan/PG-8415/2020\|EPI_ISL_687514\|2020-08 | .................... | .................... | ..................... | .................... | .................... | .................... |
| V | 4885 | hCoV-19/Japan/PG-8416/2020\|EPI_ISL_687515\|2020-08 | .................... | .................... | ..................... | .................... | .................... | .................... |
| V | 4886 | hCoV-19/Japan/PG-8417/2020\|EPI_ISL_687516\|2020-08 | .................... | .................... | ..................... | .................... | .................... | .................... |
| V | 4887 | hCoV-19/Japan/PG-8418/2020\|EPI_ISL_687517\|2020-08 | .................... | .................... | ..................... | .................... | .................... | .................... |
| V | 4888 | hCoV-19/Japan/PG-8419/2020\|EPI_ISL_687518\|2020-08 | .................... | .................... | ..................... | .................... | .................... | .................... |
| V | 4889 | hCoV-19/Japan/PG-8420/2020\|EPI_ISL_687519\|2020-08 | .................... | .................... | ..................... | .................... | .................... | .................... |
| V | 4890 | hCoV-19/Japan/PG-8421/2020\|EPI_ISL_687520\|2020-08 | .................... | .................... | ..................... | .................... | .................... | .................... |
| V | 4891 | hCoV-19/Japan/PG-8422/2020\|EPI_ISL_687521\|2020-08 | .................... | .................... | ..................... | .................... | .................... | .................... |
| V | 4892 | hCoV-19/Japan/PG-8423/2020\|EPI_ISL_687522\|2020-08 | .................... | .................... | ..................... | .................... | .................... | .................... |
| V | 4893 | hCoV-19/Japan/PG-8424/2020\|EPI_ISL_687523\|2020-09 | .................... | .................... | ..................... | .................... | .................... | .................... |
| V | 4894 | hCoV-19/Japan/PG-8425/2020\|EPI_ISL_687524\|2020-09 | .................... | .................... | ..................... | .................... | .................... | .................... |
| V | 4895 | hCoV-19/Japan/PG-8426/2020\|EPI_ISL_687525\|2020-09 | .................... | .................... | ..................... | .................... | .................... | .................... |
| V | 4896 | hCoV-19/Japan/PG-8427/2020\|EPI_ISL_687526\|2020-09 | .................... | .................... | ..................... | .................... | .................... | .................... |
| V | 4897 | hCoV-19/Japan/PG-8428/2020\|EPI_ISL_687527\|2020-09 | .................... | .................... | ..................... | .................... | .................... | .................... |
| V | 4898 | hCoV-19/Japan/PG-8429/2020\|EPI_ISL_687528\|2020-09 | .................... | .................... | ..................... | .................... | .................... | .................... |
| V | 4899 | hCoV-19/Japan/PG-8430/2020\|EPI_ISL_687529\|2020-09 | .................... | .................... | ..................... | .................... | .................... | .................... |
| V | 4900 | hCoV-19/Japan/PG-8431/2020\|EPI_ISL_687530\|2020-09 | .................... | .................... | ..................... | .................... | .................... | .................... |
| V | 4901 | hCoV-19/Japan/PG-8432/2020\|EPI_ISL_687531\|2020-09 | .................... | .................... | ..................... | .................... | .................... | .................... |
| V | 4902 | hCoV-19/Japan/PG-8433/2020\|EPI_ISL_687532\|2020-09 | .................... | .................... | ..................... | .................... | .................... | .................... |
| V | 4903 | hCoV-19/Japan/PG-8434/2020\|EPI_ISL_687533\|2020-09 | .................... | .................... | ..................... | .................... | .................... | .................... |
| V | 4904 | hCoV-19/Japan/PG-8435/2020\|EPI_ISL_687534\|2020-09 | .................... | .................... | ..................... | .................... | .................... | .................... |
| V | 4905 | hCoV-19/Japan/PG-8436/2020\|EPI_ISL_687535\|2020-09 | .................... | .................... | ..................... | .................... | .................... | .................... |
| V | 4906 | hCoV-19/Japan/PG-8437/2020\|EPI_ISL_687536\|2020-09 | .................... | .................... | ..................... | .................... | .................... | .................... |
| V | 4907 | hCoV-19/Japan/PG-8438/2020\|EPI_ISL_687537\|2020-09 | .................... | .................... | ..................... | .................... | .................... | .................... |
| V | 4908 | hCoV-19/Japan/PG-8439/2020\|EPI_ISL_687538\|2020-09 | .................... | .................... | ..................... | .................... | .................... | .................... |
| V | 4909 | hCoV-19/Japan/PG-8440/2020\|EPI_ISL_687539\|2020-09 | .................... | .................... | ..................... | .................... | .................... | .................... |
| V | 4910 | hCoV-19/Japan/PG-8441/2020\|EPI_ISL_687540\|2020-09 | .................... | .................... | ..................... | .................... | .................... | .................... |
| V | 4911 | hCoV-19/Japan/PG-8442/2020\|EPI_ISL_687541\|2020-09 | .................... | .................... | ..................... | .................... | .................... | .................... |
| V | 4912 | hCoV-19/Japan/PG-8443/2020\|EPI_ISL_687542\|2020-09 | .................... | .................... | ..................... | .................... | .................... | .................... |
| V | 4913 | hCoV-19/Japan/PG-8444/2020\|EPI_ISL_687543\|2020-09 | .................... | .................... | ..................... | .................... | .................... | .................... |
| V | 4914 | hCoV-19/Japan/PG-8445/2020\|EPI_ISL_687544\|2020-09 | .................... | .................... | ..................... | .................... | .................... | .................... |
| V | 4915 | hCoV-19/Japan/PG-8446/2020\|EPI_ISL_687545\|2020-09 | .................... | .................... | ..................... | .................... | .................... | .................... |
| V | 4916 | hCoV-19/Japan/PG-6892/2020\|EPI_ISL_688000\|2020 | .................... | .................... | ..................... | .................... | .................... | .................... |
| V | 4917 | hCoV-19/Japan/PG-6893/2020\|EPI_ISL_688001\|2020-08 | .................... | .................... | ..................... | .................... | .................... | .................... |
| V | 4918 | hCoV-19/Japan/PG-6894/2020\|EPI_ISL_688002\|2020-08 | .................... | .................... | ..................... | .................... | .................... | .................... |
| V | 4919 | hCoV-19/Japan/PG-6895/2020\|EPI_ISL_688003\|2020-08 | .................... | .................... | ..................... | .................... | .................... | .................... |
| V | 4920 | hCoV-19/Japan/PG-6896/2020\|EPI_ISL_688004\|2020-08 | .................... | .................... | ..................... | .................... | .................... | .................... |
| V | 4921 | hCoV-19/Japan/PG-6897/2020\|EPI_ISL_688005\|2020-08 | .................... | .................... | ..................... | .................... | .................... | .................... |
| V | 4922 | hCoV-19/Japan/PG-6898/2020\|EPI_ISL_688006\|2020-08 | .................... | .................... | ..................... | .................... | .................... | .................... |
| V | 4923 | hCoV-19/Japan/PG-6899/2020\|EPI_ISL_688007\|2020-09 | .................... | .................... | ..................... | .................... | .................... | .................... |
| V | 4924 | hCoV-19/Japan/PG-6900/2020\|EPI_ISL_688008\|2020-09 | .................... | .................... | ..................... | .................... | .................... | .................... |
| V | 4925 | hCoV-19/Japan/PG-6901/2020\|EPI_ISL_688009\|2020-09 | .................... | .................... | ..................... | .................... | .................... | .................... |
| V | 4926 | hCoV-19/Japan/PG-6902/2020\|EPI_ISL_688010\|2020-09 | .................... | .................... | ..................... | .................... | .................... | .................... |
| V | 4927 | hCoV-19/Japan/PG-6903/2020\|EPI_ISL_688011\|2020-09 | .................... | .................... | ..................... | .................... | .................... | .................... |
| V | 4928 | hCoV-19/Japan/PG-6904/2020\|EPI_ISL_688012\|2020-09 | .................... | .................... | ..................... | .................... | .................... | .................... |
| V | 4929 | hCoV-19/Japan/PG-6905/2020\|EPI_ISL_688013\|2020-09 | .................... | .................... | ..................... | .................... | .................... | .................... |
| V | 4930 | hCoV-19/Japan/PG-6906/2020\|EPI_ISL_688014\|2020-09 | .................... | .................... | ..................... | .................... | .................... | .................... |
| V | 4931 | hCoV-19/Japan/PG-6907/2020\|EPI_ISL_688015\|2020-09 | .................... | .................... | ..................... | .................... | .................... | .................... |
| V | 4932 | hCoV-19/Japan/PG-6908/2020\|EPI_ISL_688016\|2020-09 | .................... | .................... | ..................... | .................... | .................... | .................... |
| V | 4933 | hCoV-19/Japan/PG-6909/2020\|EPI_ISL_688017\|2020-09 | .................... | .................... | ..................... | .................... | .................... | .................... |
| V | 4934 | hCoV-19/Japan/PG-6910/2020\|EPI_ISL_688018\|2020-09 | .................... | .................... | ..................... | .................... | .................... | .................... |
| V | 4935 | hCoV-19/Japan/PG-6911/2020\|EPI_ISL_688019\|2020-09 | .................... | .................... | ..................... | .................... | .................... | .................... |
| V | 4936 | hCoV-19/Japan/PG-6912/2020\|EPI_ISL_688020\|2020-09 | .................... | .................... | ..................... | .................... | .................... | .................... |
| V | 4937 | hCoV-19/Japan/PG-5718/2020\|EPI_ISL_688840\|2020-07 | .................... | .................... | ..................... | .................... | .................... | .................... |
| V | 4938 | hCoV-19/Japan/PG-5719/2020\|EPI_ISL_688841\|2020-07 | .................... | .................... | ..................... | .................... | .................... | .................... |
| V | 4939 | hCoV-19/Japan/PG-5720/2020\|EPI_ISL_688842\|2020-07 | .................... | .................... | ..................... | .................... | .................... | .................... |
| V | 4940 | hCoV-19/Japan/PG-5721/2020\|EPI_ISL_688843\|2020-07 | .................... | .................... | ..................... | .................... | .................... | .................... |
| V | 4941 | hCoV-19/Japan/PG-5722/2020\|EPI_ISL_688844\|2020-07 | .................... | .................... | ..................... | .................... | .................... | .................... |
| V | 4942 | hCoV-19/Japan/PG-5723/2020\|EPI_ISL_688845\|2020-07 | .................... | .................... | ..................... | .................... | .................... | .................... |
| V | 4943 | hCoV-19/Japan/PG-5724/2020\|EPI_ISL_688846\|2020-07 | .................... | .................... | ..................... | .................... | .................... | .................... |
| V | 4944 | hCoV-19/Japan/PG-5725/2020\|EPI_ISL_688847\|2020-07 | .................... | .................... | ..................... | .................... | .................... | .................... |
| V | 4945 | hCoV-19/Japan/PG-5726/2020\|EPI_ISL_688848\|2020-07 | .................... | .................... | ..................... | .................... | .................... | .................... |
| V | 4946 | hCoV-19/Japan/PG-5727/2020\|EPI_ISL_688849\|2020-07 | .................... | .................... | ..................... | .................... | .................... | .................... |
| V | 4947 | hCoV-19/Japan/PG-5728/2020\|EPI_ISL_688850\|2020-07 | .................... | .................... | ..................... | .................... | .................... | .................... |
| V | 4948 | hCoV-19/Japan/PG-5729/2020\|EPI_ISL_688851\|2020-07 | .................... | .................... | ..................... | .................... | .................... | .................... |
| V | 4949 | hCoV-19/Japan/PG-5730/2020\|EPI_ISL_688852\|2020-07 | .................... | .................... | ..................... | .................... | .................... | .................... |
| V | 4950 | hCoV-19/Japan/PG-5731/2020\|EPI_ISL_688853\|2020-07 | .................... | .................... | ..................... | .................... | .................... | .................... |
| V | 4951 | hCoV-19/Japan/PG-5732/2020\|EPI_ISL_688854\|2020-07 | .................... | .................... | ..................... | .................... | .................... | .................... |
| V | 4952 | hCoV-19/Japan/PG-5735/2020\|EPI_ISL_688855\|2020-08 | .................... | .................... | ..................... | .................... | .................... | .................... |
| V | 4953 | hCoV-19/Japan/PG-5736/2020\|EPI_ISL_688856\|2020-08 | .................... | .................... | ..................... | .................... | .................... | .................... |
| V | 4954 | hCoV-19/Japan/PG-5737/2020\|EPI_ISL_688857\|2020-08 | .................... | .................... | ..................... | .................... | .................... | .................... |
| V | 4955 | hCoV-19/Japan/PG-5738/2020\|EPI_ISL_688858\|2020-08 | .................... | .................... | ..................... | .................... | .................... | .................... |
| V | 4956 | hCoV-19/Japan/PG-5739/2020\|EPI_ISL_688859\|2020-08 | .................... | .................... | ..................... | .................... | .................... | .................... |
| V | 4957 | hCoV-19/Japan/PG-5740/2020\|EPI_ISL_688860\|2020-08 | .................... | .................... | ..................... | .................... | .................... | .................... |
| V | 4958 | hCoV-19/Japan/PG-5741/2020\|EPI_ISL_688861\|2020-08 | .................... | .................... | ..................... | .................... | .................... | .................... |
| V | 4959 | hCoV-19/Japan/PG-5742/2020\|EPI_ISL_688862\|2020-08 | .................... | .................... | ..................... | .................... | .................... | .................... |
| V | 4960 | hCoV-19/Japan/PG-5743/2020\|EPI_ISL_688863\|2020-08 | .................... | .................... | ..................... | .................... | .................... | .................... |
| V | 4961 | hCoV-19/Japan/PG-5745/2020\|EPI_ISL_688864\|2020-08 | .................... | .................... | ..................... | .................... | .................... | .................... |

| | | | | | | | | |
|---|---|---|---|---|---|---|---|---|
| V | 4962 | hCoV-19/Japan/PG-5746/2020\|EPI_ISL_688865\|2020-08 | .................... | .................... | ..................... | .................... | .................... | .................... |
| V | 4963 | hCoV-19/Japan/PG-5747/2020\|EPI_ISL_688866\|2020-08 | .................... | .................... | ..................... | .................... | .................... | .................... |
| V | 4964 | hCoV-19/Japan/PG-5748/2020\|EPI_ISL_688867\|2020-08 | .................... | .................... | ..................... | .................... | .................... | .................... |
| V | 4965 | hCoV-19/Japan/PG-5749/2020\|EPI_ISL_688868\|2020-08 | .................... | .................... | ..................... | .................... | .................... | .................... |
| V | 4966 | hCoV-19/Japan/PG-5750/2020\|EPI_ISL_688869\|2020-08 | .................... | .................... | ..................... | .................... | .................... | .................... |
| V | 4967 | hCoV-19/Japan/PG-5751/2020\|EPI_ISL_688870\|2020-08 | .................... | .................... | ..................... | .................... | .................... | .................... |
| V | 4969 | hCoV-19/Japan/PG-5753/2020\|EPI_ISL_688872\|2020-08 | .................... | .................... | ..................... | .................... | .................... | .................... |
| V | 4970 | hCoV-19/Japan/PG-5754/2020\|EPI_ISL_688873\|2020-08 | .................... | .................... | ..................... | .................... | .................... | .................... |
| V | 4971 | hCoV-19/Japan/PG-5755/2020\|EPI_ISL_688874\|2020-08 | .................... | .................... | ..................... | .................... | .................... | .................... |
| V | 4972 | hCoV-19/Japan/PG-5756/2020\|EPI_ISL_688875\|2020-08 | .................... | .................... | ..................... | .................... | .................... | .................... |
| V | 4973 | hCoV-19/Japan/PG-5757/2020\|EPI_ISL_688876\|2020-08 | .................... | .................... | ..................... | .................... | .................... | .................... |
| V | 4974 | hCoV-19/Japan/PG-6216/2020\|EPI_ISL_689328\|2020-08 | .................... | .................... | ..................... | .................... | .................... | .................... |
| V | 4975 | hCoV-19/Japan/PG-6217/2020\|EPI_ISL_689329\|2020-08 | .................... | .................... | ..................... | .................... | .................... | .................... |
| V | 4976 | hCoV-19/Japan/PG-6218/2020\|EPI_ISL_689330\|2020-08 | .................... | .................... | ..................... | .................... | .................... | .................... |
| V | 4977 | hCoV-19/Japan/PG-6219/2020\|EPI_ISL_689331\|2020-08 | .................... | .................... | ..................... | .................... | .................... | .................... |
| V | 4978 | hCoV-19/Japan/PG-6220/2020\|EPI_ISL_689332\|2020-08 | .................... | .................... | ..................... | .................... | .................... | .................... |
| V | 4979 | hCoV-19/Japan/PG-6221/2020\|EPI_ISL_689333\|2020-08 | .................... | .................... | ..................... | .................... | .................... | .................... |
| V | 4980 | hCoV-19/Japan/PG-6222/2020\|EPI_ISL_689334\|2020-08 | .................... | .................... | ..................... | .................... | .................... | .................... |
| V | 4981 | hCoV-19/Japan/PG-6223/2020\|EPI_ISL_689335\|2020-08 | .................... | .................... | ..................... | .................... | .................... | .................... |
| V | 4982 | hCoV-19/Japan/PG-6224/2020\|EPI_ISL_689336\|2020-08 | .................... | .................... | ..................... | .................... | .................... | .................... |
| V | 4983 | hCoV-19/Japan/PG-6225/2020\|EPI_ISL_689337\|2020-08 | .................... | .................... | ..................... | .................... | .................... | .................... |
| V | 4984 | hCoV-19/Japan/PG-6226/2020\|EPI_ISL_689338\|2020-08 | .................... | .................... | ..................... | .................... | .................... | .................... |
| V | 4985 | hCoV-19/Japan/PG-6227/2020\|EPI_ISL_689339\|2020-08 | .................... | .................... | ..................... | .................... | .................... | .................... |
| V | 4986 | hCoV-19/Japan/PG-6228/2020\|EPI_ISL_689340\|2020-08 | .................... | .................... | ..................... | .................... | .................... | .................... |
| V | 4987 | hCoV-19/Japan/PG-6229/2020\|EPI_ISL_689341\|2020-08 | .................... | .................... | ..................... | .................... | .................... | .................... |
| V | 4988 | hCoV-19/Japan/PG-6230/2020\|EPI_ISL_689342\|2020-08 | .................... | .................... | ..................... | .................... | .................... | .................... |
| V | 4989 | hCoV-19/Japan/PG-6231/2020\|EPI_ISL_689343\|2020-08 | .................... | .................... | ..................... | .................... | .................... | .................... |
| V | 4990 | hCoV-19/Japan/PG-6232/2020\|EPI_ISL_689344\|2020-08 | .................... | .................... | ..................... | .................... | .................... | .................... |
| V | 4991 | hCoV-19/Japan/PG-6233/2020\|EPI_ISL_689345\|2020-08 | .................... | .................... | ..................... | .................... | .................... | .................... |
| V | 4992 | hCoV-19/Japan/PG-6234/2020\|EPI_ISL_689346\|2020-08 | .................... | .................... | ..................... | .................... | .................... | .................... |
| V | 4993 | hCoV-19/Japan/PG-6235/2020\|EPI_ISL_689347\|2020-08 | .................... | .................... | ..................... | .................... | .................... | .................... |
| V | 4994 | hCoV-19/Japan/PG-6236/2020\|EPI_ISL_689348\|2020-08 | .................... | .................... | ..................... | .................... | .................... | .................... |
| V | 4995 | hCoV-19/Japan/PG-6237/2020\|EPI_ISL_689349\|2020-08 | .................... | .................... | ..................... | .................... | .................... | .................... |
| V | 4996 | hCoV-19/Japan/PG-6238/2020\|EPI_ISL_689350\|2020-08 | .................... | .................... | ..................... | .................... | .................... | .................... |
| V | 4997 | hCoV-19/Japan/PG-5197/2020\|EPI_ISL_690300\|2020-08 | .................... | .................... | ..................... | .................... | .................... | .................... |
| V | 4998 | hCoV-19/Japan/PG-5198/2020\|EPI_ISL_690301\|2020-08 | .................... | .................... | ..................... | .................... | .................... | .................... |
| V | 4999 | hCoV-19/Japan/PG-5199/2020\|EPI_ISL_690302\|2020-08 | .................... | .................... | ..................... | .................... | .................... | .................... |
| V | 5000 | hCoV-19/Japan/PG-5200/2020\|EPI_ISL_690303\|2020-08 | .................... | .................... | ..................... | .................... | .................... | .................... |
| V | 5001 | hCoV-19/Japan/PG-5201/2020\|EPI_ISL_690304\|2020-08 | .................... | .................... | ..................... | .................... | .................... | .................... |
| V | 5003 | hCoV-19/Japan/PG-5203/2020\|EPI_ISL_690306\|2020-08 | .................... | .................... | ..................... | .................... | .................... | .................... |
| V | 5004 | hCoV-19/Japan/PG-5204/2020\|EPI_ISL_690307\|2020-08 | .................... | .................... | ..................... | .................... | .................... | .................... |
| V | 5005 | hCoV-19/Japan/PG-5205/2020\|EPI_ISL_690308\|2020-08 | .................... | .................... | ..................... | .................... | .................... | .................... |
| V | 5006 | hCoV-19/Japan/PG-5206/2020\|EPI_ISL_690309\|2020-08 | .................... | .................... | ..................... | .................... | .................... | .................... |
| V | 5007 | hCoV-19/Japan/PG-3501/2020\|EPI_ISL_691610\|2020-07 | .................... | .................... | ..................... | .................... | .................... | .................... |
| V | 5008 | hCoV-19/Japan/PG-3502/2020\|EPI_ISL_691611\|2020-07 | .................... | .................... | ..................... | .................... | .................... | .................... |
| V | 5009 | hCoV-19/Japan/PG-3747/2020\|EPI_ISL_692000\|2020-04 | .................... | .................... | ..................... | .................... | .................... | .................... |
| V | 5010 | hCoV-19/Japan/PG-3748/2020\|EPI_ISL_692001\|2020-04 | .................... | .................... | ..................... | .................... | .................... | .................... |
| V | 5011 | hCoV-19/Japan/PG-3749/2020\|EPI_ISL_692002\|2020-04 | .................... | .................... | ..................... | .................... | .................... | .................... |
| V | 5012 | hCoV-19/Japan/PG-3750/2020\|EPI_ISL_692003\|2020-04 | .................... | .................... | ..................... | .................... | .................... | .................... |
| V | 5013 | hCoV-19/Japan/PG-3751/2020\|EPI_ISL_692004\|2020-04 | .................... | .................... | ..................... | .................... | .................... | .................... |
| V | 5014 | hCoV-19/Japan/PG-3752/2020\|EPI_ISL_692005\|2020-04 | .................... | .................... | ..................... | .................... | .................... | .................... |
| V | 5015 | hCoV-19/Japan/PG-3753/2020\|EPI_ISL_692006\|2020-04 | .................... | .................... | ..................... | .................... | .................... | .................... |
| V | 5016 | hCoV-19/Japan/PG-3754/2020\|EPI_ISL_692007\|2020-04 | .................... | .................... | ..................... | .................... | .................... | .................... |
| V | 5017 | hCoV-19/Japan/PG-3755/2020\|EPI_ISL_692008\|2020-04 | .................... | .................... | ..................... | .................... | .................... | .................... |
| V | 5018 | hCoV-19/Japan/PG-3756/2020\|EPI_ISL_692009\|2020-04 | .................... | .................... | ..................... | .................... | .................... | .................... |
| V | 5019 | hCoV-19/Japan/PG-3757/2020\|EPI_ISL_692010\|2020-04 | .................... | .................... | ..................... | .................... | .................... | .................... |
| V | 5020 | hCoV-19/Japan/PG-3805/2020\|EPI_ISL_692056\|2020-07 | .................... | .................... | ..................... | .................... | .................... | .................... |
| V | 5021 | hCoV-19/Japan/PG-3806/2020\|EPI_ISL_692057\|2020-07 | .................... | .................... | ..................... | .................... | .................... | .................... |
| V | 5022 | hCoV-19/Japan/PG-3807/2020\|EPI_ISL_692058\|2020-07 | .................... | .................... | ..................... | .................... | .................... | .................... |
| V | 5023 | hCoV-19/Japan/PG-3808/2020\|EPI_ISL_692059\|2020-05 | .................... | .................... | ..................... | .................... | .................... | .................... |
| V | 5024 | hCoV-19/Japan/PG-3809/2020\|EPI_ISL_692060\|2020-05 | .................... | .................... | ..................... | .................... | .................... | .................... |
| V | 5025 | hCoV-19/Japan/PG-3810/2020\|EPI_ISL_692061\|2020-06 | .................... | .................... | ..................... | .................... | .................... | .................... |
| V | 5026 | hCoV-19/Japan/PG-3811/2020\|EPI_ISL_692062\|2020-06 | .................... | .................... | ..................... | .................... | .................... | .................... |
| V | 5027 | hCoV-19/Japan/PG-3812/2020\|EPI_ISL_692063\|2020-06 | .................... | .................... | ..................... | .................... | .................... | .................... |
| V | 5028 | hCoV-19/Japan/PG-3813/2020\|EPI_ISL_692064\|2020-06 | .................... | .................... | ..................... | .................... | .................... | .................... |
| V | 5029 | hCoV-19/Japan/PG-3814/2020\|EPI_ISL_692065\|2020-06 | .................... | .................... | ..................... | .................... | .................... | .................... |
| V | 5030 | hCoV-19/Japan/PG-3815/2020\|EPI_ISL_692066\|2020-06 | .................... | .................... | ..................... | .................... | .................... | .................... |
| V | 5031 | hCoV-19/Japan/PG-3816/2020\|EPI_ISL_692067\|2020-06 | .................... | .................... | ..................... | .................... | .................... | .................... |
| V | 5032 | hCoV-19/Japan/PG-3817/2020\|EPI_ISL_692068\|2020-06 | .................... | .................... | ..................... | .................... | .................... | .................... |
| V | 5033 | hCoV-19/Japan/PG-3818/2020\|EPI_ISL_692069\|2020-06 | .................... | .................... | ..................... | .................... | .................... | .................... |
| V | 5034 | hCoV-19/Japan/PG-3819/2020\|EPI_ISL_692070\|2020-06 | .................... | .................... | ..................... | .................... | .................... | .................... |
| V | 5035 | hCoV-19/Japan/PG-3820/2020\|EPI_ISL_692071\|2020-06 | .................... | .................... | ..................... | .................... | .................... | .................... |
| V | 5036 | hCoV-19/Japan/PG-3821/2020\|EPI_ISL_692072\|2020-07 | .................... | .................... | ..................... | .................... | .................... | .................... |
| V | 5037 | hCoV-19/Japan/PG-3822/2020\|EPI_ISL_692073\|2020-07 | .................... | .................... | ..................... | .................... | .................... | .................... |
| V | 5038 | hCoV-19/Japan/PG-3823/2020\|EPI_ISL_692074\|2020-07 | .................... | .................... | ..................... | .................... | .................... | .................... |
| V | 5039 | hCoV-19/Japan/PG-3824/2020\|EPI_ISL_692075\|2020-07 | .................... | .................... | ..................... | .................... | .................... | .................... |
| V | 5040 | hCoV-19/Japan/PG-3825/2020\|EPI_ISL_692076\|2020-07 | .................... | .................... | ..................... | .................... | .................... | .................... |
| V | 5041 | hCoV-19/Japan/PG-3826/2020\|EPI_ISL_692077\|2020-07 | .................... | .................... | ..................... | .................... | .................... | .................... |
| V | 5042 | hCoV-19/Japan/PG-3827/2020\|EPI_ISL_692078\|2020-07 | .................... | .................... | ..................... | .................... | .................... | .................... |
| V | 5043 | hCoV-19/Japan/PG-3828/2020\|EPI_ISL_692079\|2020-07 | .................... | .................... | ..................... | .................... | .................... | .................... |

| | | | | | | | |
|---|---|---|---|---|---|---|---|
| V | 5044 hCoV-19/Japan/PG-3829/2020\|EPI_ISL_692080\|2020-07 | ···················· | ···················· | ····················· | ···················· | ···················· | ···················· |
| V | 5045 hCoV-19/Japan/PG-3830/2020\|EPI_ISL_692081\|2020-07 | ···················· | ···················· | ····················· | ···················· | ···················· | ···················· |
| V | 5046 hCoV-19/Japan/PG-3831/2020\|EPI_ISL_692082\|2020-07 | ···················· | ···················· | ····················· | ···················· | ···················· | ···················· |
| V | 5047 hCoV-19/Japan/PG-3832/2020\|EPI_ISL_692083\|2020-07 | ···················· | ···················· | ····················· | ···················· | ···················· | ···················· |
| V | 5048 hCoV-19/Japan/PG-3833/2020\|EPI_ISL_692084\|2020-07 | ···················· | ···················· | ····················· | ···················· | ···················· | ···················· |
| V | 5049 hCoV-19/Japan/PG-3834/2020\|EPI_ISL_692085\|2020-07 | ···················· | ···················· | ····················· | ···················· | ···················· | ···················· |
| V | 5050 hCoV-19/Japan/PG-3835/2020\|EPI_ISL_692086\|2020-07 | ···················· | ···················· | ····················· | ···················· | ···················· | ···················· |
| V | 5051 hCoV-19/Japan/PG-3836/2020\|EPI_ISL_692087\|2020-07 | ···················· | ···················· | ····················· | ···················· | ···················· | ···················· |
| V | 5052 hCoV-19/Japan/PG-3837/2020\|EPI_ISL_692088\|2020-07 | ···················· | ···················· | ····················· | ···················· | ···················· | ···················· |
| V | 5053 hCoV-19/Japan/PG-3838/2020\|EPI_ISL_692089\|2020-07 | ···················· | ···················· | ····················· | ···················· | ···················· | ···················· |
| V | 5054 hCoV-19/Japan/PG-3839/2020\|EPI_ISL_692090\|2020-07 | ···················· | ···················· | ····················· | ···················· | ···················· | ···················· |
| V | 5055 hCoV-19/Japan/PG-3840/2020\|EPI_ISL_692091\|2020-07 | ···················· | ···················· | ····················· | ···················· | ···················· | ···················· |
| V | 5056 hCoV-19/Japan/PG-3841/2020\|EPI_ISL_692092\|2020-07 | ···················· | ···················· | ····················· | ···················· | ···················· | ···················· |
| V | 5057 hCoV-19/Japan/PG-3842/2020\|EPI_ISL_692093\|2020-07 | ···················· | ···················· | ····················· | ···················· | ···················· | ···················· |
| V | 5058 hCoV-19/Japan/PG-3843/2020\|EPI_ISL_692094\|2020-07 | ···················· | ···················· | ····················· | ···················· | ···················· | ···················· |
| V | 5059 hCoV-19/Japan/PG-3844/2020\|EPI_ISL_692095\|2020-07 | ···················· | ···················· | ····················· | ···················· | ···················· | ···················· |
| V | 5060 hCoV-19/Japan/PG-3845/2020\|EPI_ISL_692096\|2020-07 | ···················· | ···················· | ····················· | ···················· | ···················· | ···················· |
| V | 5061 hCoV-19/Japan/PG-3846/2020\|EPI_ISL_692097\|2020-07 | ···················· | ···················· | ····················· | ···················· | ···················· | ···················· |
| V | 5062 hCoV-19/Japan/PG-3847/2020\|EPI_ISL_692098\|2020-07 | ···················· | ···················· | ····················· | ···················· | ···················· | ···················· |
| V | 5063 hCoV-19/Japan/PG-3848/2020\|EPI_ISL_692099\|2020-07 | ···················· | ···················· | ····················· | ···················· | ···················· | ···················· |
| V | 5064 hCoV-19/USA/MA-MASPHL-00732/2020\|EPI_ISL_692940\|20 | ···················· | ···················· | ····················· | ···················· | ···················· | ···················· |
| V | 5065 hCoV-19/USA/MA-MASPHL-00731/2020\|EPI_ISL_692941\|20 | ···················· | ···················· | ····················· | ···················· | ···················· | ···················· |
| V | 5066 hCoV-19/USA/MA-MASPHL-00730/2020\|EPI_ISL_692942\|20 | ···················· | ···················· | ····················· | ···················· | ···················· | ···················· |
| V | 5067 hCoV-19/USA/MA-MASPHL-00729/2020\|EPI_ISL_692943\|20 | ···················· | ···················· | ····················· | ···················· | ···················· | ···················· |
| V | 5068 hCoV-19/USA/MA-MASPHL-00697/2020\|EPI_ISL_692950\|20 | ···················· | ···················· | ····················· | ···················· | ···················· | ···················· |
| V | 5069 hCoV-19/USA/MA-MASPHL-00695/2020\|EPI_ISL_692952\|20 | ···················· | ···················· | ····················· | ···················· | ···················· | ···················· |
| V | 5070 hCoV-19/USA/MA-MASPHL-00694/2020\|EPI_ISL_692953\|20 | ···················· | ···················· | ····················· | ···················· | ···················· | ···················· |
| V | 5071 hCoV-19/USA/MA-MASPHL-00690/2020\|EPI_ISL_692956\|20 | ···················· | ···················· | ····················· | ···················· | ···················· | ···················· |
| V | 5072 hCoV-19/USA/MA-MASPHL-00689/2020\|EPI_ISL_692957\|20 | ···················· | ···················· | ····················· | ···················· | ···················· | ···················· |
| V | 5073 hCoV-19/USA/MA-MASPHL-00685/2020\|EPI_ISL_692961\|20 | ···················· | ···················· | ····················· | ···················· | ···················· | ···················· |
| V | 5074 hCoV-19/USA/MA-MASPHL-00684/2020\|EPI_ISL_692962\|20 | ···················· | ···················· | ····················· | ···················· | ···················· | ···················· |
| V | 5075 hCoV-19/USA/MA-MASPHL-00682/2020\|EPI_ISL_692963\|20 | ···················· | ···················· | ····················· | ···················· | ···················· | ···················· |
| V | 5076 hCoV-19/USA/MA-MASPHL-00681/2020\|EPI_ISL_692964\|20 | ···················· | ···················· | ····················· | ···················· | ···················· | ···················· |
| V | 5077 hCoV-19/USA/MA-MASPHL-00680/2020\|EPI_ISL_692965\|20 | ···················· | ···················· | ····················· | ···················· | ···················· | ···················· |
| V | 5078 hCoV-19/USA/MA-MASPHL-00679/2020\|EPI_ISL_692966\|20 | ···················· | ···················· | ····················· | ···················· | ···················· | ···················· |
| V | 5079 hCoV-19/USA/MA-MASPHL-00669/2020\|EPI_ISL_692968\|20 | ···················· | ···················· | ····················· | ···················· | ···················· | ···················· |
| V | 5080 hCoV-19/Taiwan/NTU31/2020\|EPI_ISL_693302\|2020-11-1 | ···················· | ·A·················· | ····················· | ···················· | ···················· | ···A················ |
| V | 5081 hCoV-19/Taiwan/NTU32/2020\|EPI_ISL_693303\|2020-11-1 | ···················· | ···················· | ····················· | ···················· | ···················· | ···················· |
| V | 5082 hCoV-19/Taiwan/NTU33/2020\|EPI_ISL_693304\|2020-11-2 | ···················· | ···················· | ····················· | ···················· | ···················· | ···················· |
| V | 5083 hCoV-19/Taiwan/NTU34/2020\|EPI_ISL_693305\|2020-11-2 | ···················· | ···················· | ····················· | ···················· | ···················· | ···················· |
| V | 5084 hCoV-19/Taiwan/NTU35/2020\|EPI_ISL_693306\|2020-11-2 | ···················· | ·A·················· | ····················· | ···················· | ···················· | ···················· |
| V | 5085 hCoV-19/Singapore/1419/2020\|EPI_ISL_693307\|2020-11 | ···················· | ·A·················· | ····················· | ···················· | ···················· | ···················· |
| V | 5086 hCoV-19/Singapore/1420/2020\|EPI_ISL_693308\|2020-11 | ···················· | ·A·················· | ····················· | ···················· | ···················· | ···A················ |
| V | 5087 hCoV-19/Singapore/1421/2020\|EPI_ISL_693309\|2020-11 | ···················· | ·A·················· | ····················· | ···················· | ···················· | ···················· |
| V | 5088 hCoV-19/Singapore/1422/2020\|EPI_ISL_693310\|2020-11 | ···················· | ·A·················· | ····················· | ···················· | ···················· | ···A················ |
| V | 5089 hCoV-19/Singapore/1423/2020\|EPI_ISL_693311\|2020-11 | ···················· | ·A·················· | ····················· | ···················· | ···················· | ···A················ |
| V | 5090 hCoV-19/Singapore/1424/2020\|EPI_ISL_693312\|2020-11 | ···················· | ···················· | ····················· | ···················· | ···················· | ···················· |
| V | 5091 hCoV-19/Singapore/1425/2020\|EPI_ISL_693313\|2020-11 | ···················· | ···················· | ····················· | ···················· | ···················· | ···················· |
| V | 5092 hCoV-19/Singapore/1426/2020\|EPI_ISL_693314\|2020-11 | ···················· | ···················· | ····················· | ···················· | ···················· | ···················· |
| V | 5093 hCoV-19/Singapore/1427/2020\|EPI_ISL_693315\|2020-11 | ···················· | ·A·················· | ····················· | ···················· | ···················· | ···················· |
| V | 5094 hCoV-19/Singapore/1428/2020\|EPI_ISL_693316\|2020-11 | ···················· | ·A·················· | ····················· | ···················· | ···················· | ···A················ |
| V | 5095 hCoV-19/Singapore/1429/2020\|EPI_ISL_693317\|2020-11 | ···················· | ···················· | ····················· | ···················· | ···················· | ···················· |
| V | 5096 hCoV-19/Singapore/1430/2020\|EPI_ISL_693318\|2020-11 | ···················· | ···················· | ····················· | ···················· | ···················· | ···················· |
| V | 5097 hCoV-19/Singapore/1431/2020\|EPI_ISL_693319\|2020-11 | ···················· | ···················· | ····················· | ···················· | ···················· | ···················· |
| V | 5098 hCoV-19/Singapore/1432/2020\|EPI_ISL_693320\|2020-11 | ···················· | ·A·················· | ····················· | ···················· | ···················· | ···A················ |
| V | 5099 hCoV-19/Singapore/1433/2020\|EPI_ISL_693321\|2020-11 | ···················· | ·A·················· | ····················· | ···················· | ···················· | ···A················ |
| V | 5100 hCoV-19/Singapore/1434/2020\|EPI_ISL_693322\|2020-11 | ···················· | ·A·················· | ····················· | ···················· | ···················· | ···················· |
| V | 5101 hCoV-19/Singapore/1435/2020\|EPI_ISL_693323\|2020-11 | ···················· | ·A·················· | ····················· | ···················· | ···················· | ···················· |
| V | 5102 hCoV-19/Singapore/1436/2020\|EPI_ISL_693324\|2020-11 | ···················· | ···················· | ····················· | ···················· | ···················· | ···················· |
| V | 5103 hCoV-19/Singapore/1437/2020\|EPI_ISL_693325\|2020-11 | ···················· | ···················· | ····················· | ···················· | ···················· | ···A················ |
| V | 5104 hCoV-19/Singapore/1438/2020\|EPI_ISL_693326\|2020-11 | ···················· | ···················· | ····················· | ···················· | ···················· | ···················· |
| V | 5105 hCoV-19/Singapore/1439/2020\|EPI_ISL_693327\|2020-11 | ···················· | ···················· | ····················· | ···················· | ···················· | ···················· |
| V | 5106 hCoV-19/Singapore/1434-2/2020\|EPI_ISL_693328\|2020- | ···················· | ·A·················· | ····················· | ···················· | ···················· | ···················· |
| V | 5107 hCoV-19/Singapore/1440/2020\|EPI_ISL_693329\|2020-11 | ···················· | ···················· | ····················· | ···················· | ···················· | ···················· |
| V | 5108 hCoV-19/Singapore/1441/2020\|EPI_ISL_693330\|2020-11 | ···················· | ·A·················· | ····················· | ···················· | ···················· | ···················· |
| V | 5109 hCoV-19/Singapore/1442/2020\|EPI_ISL_693331\|2020-11 | ···················· | ·A·················· | ····················· | ···················· | ···················· | ···················· |
| V | 5110 hCoV-19/England/ALDP-BB36B4/2020\|EPI_ISL_693332\|20 | ···················· | ···················· | ····················· | ···················· | ···················· | ···················· |
| V | 5116 hCoV-19/France/PAC02003/2020\|EPI_ISL_693387\|2020-0 | ···················· | ···················· | ····················· | ···················· | ···················· | ···················· |
| V | 5117 hCoV-19/France/PAC02004/2020\|EPI_ISL_693388\|2020-0 | ···················· | ···················· | ····················· | ···················· | ···················· | ···················· |
| V | 5118 hCoV-19/France/PAC02005/2020\|EPI_ISL_693389\|2020-0 | ···················· | ···················· | ····················· | ···················· | ···················· | ···················· |
| V | 5119 hCoV-19/France/PAC02011/2020\|EPI_ISL_693390\|2020-0 | ···················· | ···················· | ····················· | ···················· | ···················· | ···················· |
| V | 5120 hCoV-19/France/ARA-SC088/2020\|EPI_ISL_693391\|2020- | ···················· | ···················· | ····················· | ···················· | ···················· | ···················· |
| V | 5121 hCoV-19/Japan/PG-8527/2020\|EPI_ISL_686123\|2020-07 | ···················· | ···················· | ····················· | ···················· | ···················· | ···················· |
| V | 5122 hCoV-19/Japan/PG-8528/2020\|EPI_ISL_686124\|2020-07 | ···················· | ···················· | ····················· | ···················· | ···················· | ···················· |
| V | 5123 hCoV-19/Japan/PG-8529/2020\|EPI_ISL_686125\|2020-07 | ···················· | ···················· | ····················· | ···················· | ···················· | ···················· |
| V | 5124 hCoV-19/Japan/PG-8530/2020\|EPI_ISL_686126\|2020-07 | ···················· | ···················· | ····················· | ···················· | ···················· | ···················· |
| V | 5125 hCoV-19/Japan/PG-8531/2020\|EPI_ISL_686127\|2020-07 | ···················· | ···················· | ····················· | ···················· | ···················· | ···················· |
| V | 5126 hCoV-19/Japan/PG-8532/2020\|EPI_ISL_686128\|2020-07 | ···················· | ···················· | ····················· | ···················· | ···················· | ···················· |
| V | 5127 hCoV-19/Japan/PG-8533/2020\|EPI_ISL_686129\|2020-07 | ···················· | ···················· | ····················· | ···················· | ···················· | ···················· |
| V | 5128 hCoV-19/Japan/PG-8534/2020\|EPI_ISL_686130\|2020-07 | ···················· | ···················· | ····················· | ···················· | ···················· | ···················· |

| | | | | | | | | |
|---|---|---|---|---|---|---|---|---|
| V | 5129 | hCoV-19/Japan/PG-8535/2020\|EPI_ISL_686131\|2020-07 | .................... | .................... | .................... | .................... | .................... | .................... |
| V | 5130 | hCoV-19/Japan/PG-8536/2020\|EPI_ISL_686132\|2020-07 | .................... | .................... | .................... | .................... | .................... | .................... |
| V | 5131 | hCoV-19/Japan/PG-8537/2020\|EPI_ISL_686133\|2020-07 | .................... | .................... | .................... | .................... | .................... | .................... |
| V | 5132 | hCoV-19/Japan/PG-8538/2020\|EPI_ISL_686134\|2020-07 | .................... | .................... | .................... | .................... | .................... | .................... |
| V | 5133 | hCoV-19/Japan/PG-8539/2020\|EPI_ISL_686135\|2020-07 | .................... | .................... | .................... | .................... | .................... | .................... |
| V | 5134 | hCoV-19/Japan/PG-8540/2020\|EPI_ISL_686136\|2020-07 | .................... | .................... | .................... | .................... | .................... | .................... |
| V | 5135 | hCoV-19/Japan/PG-8541/2020\|EPI_ISL_686137\|2020-08 | .................... | .................... | .................... | .................... | .................... | .................... |
| V | 5136 | hCoV-19/Japan/PG-8542/2020\|EPI_ISL_686138\|2020-08 | .................... | .................... | .................... | .................... | .................... | .................... |
| V | 5137 | hCoV-19/Japan/PG-8543/2020\|EPI_ISL_686139\|2020-08 | .................... | .................... | .................... | .................... | .................... | .................... |
| V | 5138 | hCoV-19/Japan/PG-8544/2020\|EPI_ISL_686140\|2020-08 | .................... | .................... | .................... | .................... | .................... | .................... |
| V | 5139 | hCoV-19/Japan/PG-8545/2020\|EPI_ISL_686141\|2020-08 | .................... | .................... | .................... | .................... | .................... | .................... |
| V | 5140 | hCoV-19/Japan/PG-8546/2020\|EPI_ISL_686142\|2020-08 | .................... | .................... | .................... | .................... | .................... | .................... |
| V | 5141 | hCoV-19/Japan/PG-8547/2020\|EPI_ISL_686143\|2020-08 | .................... | .................... | .................... | .................... | .................... | .................... |
| V | 5142 | hCoV-19/Japan/PG-8548/2020\|EPI_ISL_686144\|2020-08 | .................... | .................... | .................... | .................... | .................... | .................... |
| V | 5143 | hCoV-19/Japan/PG-8549/2020\|EPI_ISL_686145\|2020-08 | .................... | .................... | .................... | .................... | .................... | .................... |
| V | 5144 | hCoV-19/Japan/PG-8550/2020\|EPI_ISL_686146\|2020-08 | .................... | .................... | .................... | .................... | .................... | .................... |
| V | 5145 | hCoV-19/Japan/PG-8551/2020\|EPI_ISL_686147\|2020-08 | .................... | .................... | .................... | .................... | .................... | .................... |
| V | 5146 | hCoV-19/Japan/PG-8552/2020\|EPI_ISL_686148\|2020-08 | .................... | .................... | .................... | .................... | .................... | .................... |
| V | 5147 | hCoV-19/Japan/PG-8553/2020\|EPI_ISL_686149\|2020-08 | .................... | .................... | .................... | .................... | .................... | .................... |
| V | 5148 | hCoV-19/Japan/PG-8554/2020\|EPI_ISL_686150\|2020-08 | .................... | .................... | .................... | .................... | .................... | .................... |
| V | 5149 | hCoV-19/Japan/PG-8555/2020\|EPI_ISL_686151\|2020-08 | .................... | .................... | .................... | .................... | .................... | .................... |
| V | 5150 | hCoV-19/Japan/PG-8556/2020\|EPI_ISL_686152\|2020-08 | .................... | .................... | .................... | .................... | .................... | .................... |
| V | 5151 | hCoV-19/Japan/PG-8557/2020\|EPI_ISL_686153\|2020-08 | .................... | .................... | .................... | .................... | .................... | .................... |
| V | 5152 | hCoV-19/Japan/PG-8558/2020\|EPI_ISL_686154\|2020-08 | .................... | .................... | .................... | .................... | .................... | .................... |
| V | 5153 | hCoV-19/Japan/PG-8559/2020\|EPI_ISL_686155\|2020-08 | .................... | .................... | .................... | .................... | .................... | .................... |
| V | 5154 | hCoV-19/Japan/PG-8560/2020\|EPI_ISL_686156\|2020-08 | .................... | .................... | .................... | .................... | .................... | .................... |
| V | 5155 | hCoV-19/Japan/PG-8561/2020\|EPI_ISL_686157\|2020-08 | .................... | .................... | .................... | .................... | .................... | .................... |
| V | 5156 | hCoV-19/Japan/PG-8562/2020\|EPI_ISL_686158\|2020-08 | .................... | .................... | .................... | .................... | .................... | .................... |
| V | 5157 | hCoV-19/Japan/PG-8563/2020\|EPI_ISL_686159\|2020-08 | .................... | .................... | .................... | .................... | .................... | .................... |
| V | 5158 | hCoV-19/Japan/PG-8564/2020\|EPI_ISL_686160\|2020-08 | .................... | .................... | .................... | .................... | .................... | .................... |
| V | 5159 | hCoV-19/Japan/PG-8565/2020\|EPI_ISL_686161\|2020-08 | .................... | .................... | .................... | .................... | .................... | .................... |
| V | 5160 | hCoV-19/Japan/PG-8566/2020\|EPI_ISL_686162\|2020-08 | .................... | .................... | .................... | .................... | .................... | .................... |
| V | 5161 | hCoV-19/Japan/PG-8567/2020\|EPI_ISL_686163\|2020-08 | .................... | .................... | .................... | .................... | .................... | .................... |
| V | 5162 | hCoV-19/Japan/PG-8568/2020\|EPI_ISL_686164\|2020-08 | .................... | .................... | .................... | .................... | .................... | .................... |
| V | 5163 | hCoV-19/Japan/PG-8569/2020\|EPI_ISL_686165\|2020-08 | .................... | .................... | .................... | .................... | .................... | .................... |
| V | 5164 | hCoV-19/Japan/PG-8570/2020\|EPI_ISL_686166\|2020-08 | .................... | .................... | .................... | .................... | .................... | .................... |
| V | 5165 | hCoV-19/Japan/PG-8571/2020\|EPI_ISL_686167\|2020-08 | .................... | .................... | .................... | .................... | .................... | .................... |
| V | 5166 | hCoV-19/Japan/PG-8572/2020\|EPI_ISL_686168\|2020-08 | .................... | .................... | .................... | .................... | .................... | .................... |
| V | 5167 | hCoV-19/Japan/PG-8573/2020\|EPI_ISL_686169\|2020-08 | .................... | .................... | .................... | .................... | .................... | .................... |
| V | 5168 | hCoV-19/Japan/PG-8574/2020\|EPI_ISL_686170\|2020-08 | .................... | .................... | .................... | .................... | .................... | .................... |
| V | 5169 | hCoV-19/Japan/PG-8575/2020\|EPI_ISL_686171\|2020-08 | .................... | .................... | .................... | .................... | .................... | .................... |
| V | 5170 | hCoV-19/Japan/PG-8576/2020\|EPI_ISL_686172\|2020-08 | .................... | .................... | .................... | .................... | .................... | .................... |
| V | 5171 | hCoV-19/Japan/PG-8577/2020\|EPI_ISL_686173\|2020-08 | .................... | .................... | .................... | .................... | .................... | .................... |
| V | 5172 | hCoV-19/Japan/PG-8578/2020\|EPI_ISL_686174\|2020-08 | .................... | .................... | .................... | .................... | .................... | .................... |
| V | 5173 | hCoV-19/Japan/PG-8579/2020\|EPI_ISL_686175\|2020-08 | .................... | .................... | .................... | .................... | .................... | .................... |
| V | 5174 | hCoV-19/Japan/PG-8580/2020\|EPI_ISL_686176\|2020-08 | .................... | .................... | .................... | .................... | .................... | .................... |
| V | 5175 | hCoV-19/Japan/PG-8581/2020\|EPI_ISL_686177\|2020-08 | .................... | .................... | .................... | .................... | .................... | .................... |
| V | 5176 | hCoV-19/Japan/PG-8582/2020\|EPI_ISL_686178\|2020-08 | .................... | .................... | .................... | .................... | .................... | .................... |
| V | 5177 | hCoV-19/Japan/PG-8583/2020\|EPI_ISL_686179\|2020-08 | .................... | .................... | .................... | .................... | .................... | .................... |
| V | 5178 | hCoV-19/Japan/PG-8355/2020\|EPI_ISL_687454\|2020-08 | .................... | .................... | .................... | .................... | .................... | .................... |
| V | 5179 | hCoV-19/Japan/PG-8356/2020\|EPI_ISL_687455\|2020-08 | .................... | .................... | .................... | .................... | .................... | .................... |
| V | 5180 | hCoV-19/Japan/PG-8357/2020\|EPI_ISL_687456\|2020-08 | .................... | .................... | .................... | .................... | .................... | .................... |
| V | 5181 | hCoV-19/Japan/PG-8358/2020\|EPI_ISL_687457\|2020-09 | .................... | .................... | .................... | .................... | .................... | .................... |
| V | 5182 | hCoV-19/Japan/PG-8359/2020\|EPI_ISL_687458\|2020-09 | .................... | .................... | .................... | .................... | .................... | .................... |
| V | 5183 | hCoV-19/Japan/PG-8360/2020\|EPI_ISL_687459\|2020-06 | .................... | .................... | .................... | .................... | .................... | .................... |
| V | 5184 | hCoV-19/Japan/PG-8361/2020\|EPI_ISL_687460\|2020-07 | .................... | .................... | .................... | .................... | .................... | .................... |
| V | 5185 | hCoV-19/Japan/PG-8362/2020\|EPI_ISL_687461\|2020-07 | .................... | .................... | .................... | .................... | .................... | .................... |
| V | 5186 | hCoV-19/Japan/PG-8363/2020\|EPI_ISL_687462\|2020-07 | .................... | .................... | .................... | .................... | .................... | .................... |
| V | 5187 | hCoV-19/Japan/PG-8364/2020\|EPI_ISL_687463\|2020-07 | .................... | .................... | .................... | .................... | .................... | .................... |
| V | 5189 | hCoV-19/Japan/PG-8366/2020\|EPI_ISL_687465\|2020-07 | .................... | .................... | .................... | .................... | .................... | .................... |
| V | 5190 | hCoV-19/Japan/PG-8367/2020\|EPI_ISL_687466\|2020-07 | .................... | .................... | .................... | .................... | .................... | .................... |
| V | 5191 | hCoV-19/Japan/PG-8368/2020\|EPI_ISL_687467\|2020-07 | .................... | .................... | .................... | .................... | .................... | .................... |
| V | 5192 | hCoV-19/Japan/PG-8369/2020\|EPI_ISL_687468\|2020-07 | .................... | .................... | .................... | .................... | .................... | .................... |
| V | 5193 | hCoV-19/Japan/PG-8370/2020\|EPI_ISL_687469\|2020-07 | .................... | .................... | .................... | .................... | .................... | .................... |
| V | 5194 | hCoV-19/Japan/PG-8371/2020\|EPI_ISL_687470\|2020-07 | .................... | .................... | .................... | .................... | .................... | .................... |
| V | 5195 | hCoV-19/Japan/PG-8372/2020\|EPI_ISL_687471\|2020-07 | .................... | .................... | .................... | .................... | .................... | .................... |
| V | 5196 | hCoV-19/Japan/PG-8373/2020\|EPI_ISL_687472\|2020-07 | .................... | .................... | .................... | .................... | .................... | .................... |
| V | 5198 | hCoV-19/Japan/PG-8375/2020\|EPI_ISL_687474\|2020-07 | .................... | .................... | .................... | .................... | .................... | .................... |
| V | 5199 | hCoV-19/Japan/PG-8376/2020\|EPI_ISL_687475\|2020-07 | .................... | .................... | .................... | .................... | .................... | .................... |
| V | 5200 | hCoV-19/Japan/PG-8377/2020\|EPI_ISL_687476\|2020-07 | .................... | .................... | .................... | .................... | .................... | .................... |
| V | 5201 | hCoV-19/Japan/PG-8378/2020\|EPI_ISL_687477\|2020-07 | .................... | .................... | .................... | .................... | .................... | .................... |
| V | 5202 | hCoV-19/Japan/PG-8379/2020\|EPI_ISL_687478\|2020-07 | .................... | .................... | .................... | .................... | .................... | .................... |
| V | 5203 | hCoV-19/Japan/PG-8380/2020\|EPI_ISL_687479\|2020-07 | .................... | .................... | .................... | .................... | .................... | .................... |
| V | 5204 | hCoV-19/Japan/PG-8381/2020\|EPI_ISL_687480\|2020-07 | .................... | .................... | .................... | .................... | .................... | .................... |
| V | 5205 | hCoV-19/Japan/PG-8382/2020\|EPI_ISL_687481\|2020-07 | .................... | .................... | .................... | .................... | .................... | .................... |
| V | 5206 | hCoV-19/Japan/PG-8383/2020\|EPI_ISL_687482\|2020-07 | .................... | .................... | .................... | .................... | .................... | .................... |
| V | 5207 | hCoV-19/Japan/PG-8384/2020\|EPI_ISL_687483\|2020-07 | .................... | .................... | .................... | .................... | .................... | .................... |
| V | 5208 | hCoV-19/Japan/PG-8385/2020\|EPI_ISL_687484\|2020-07 | .................... | .................... | .................... | .................... | .................... | .................... |
| V | 5209 | hCoV-19/Japan/PG-8386/2020\|EPI_ISL_687485\|2020-07 | .................... | .................... | .................... | .................... | .................... | .................... |
| V | 5210 | hCoV-19/Japan/PG-8387/2020\|EPI_ISL_687486\|2020-07 | .................... | .................... | .................... | .................... | .................... | .................... |

| | | | | | | | |
|---|---|---|---|---|---|---|---|
| V | 5211 hCoV-19/Japan/PG-8388/2020\|EPI_ISL_687487\|2020-07 | .................... | .................... | .................... | .................... | .................... | .................... |
| V | 5212 hCoV-19/Japan/PG-8389/2020\|EPI_ISL_687488\|2020-07 | .................... | .................... | .................... | .................... | .................... | .................... |
| V | 5213 hCoV-19/Japan/PG-8390/2020\|EPI_ISL_687489\|2020-07 | .................... | .................... | .................... | .................... | .................... | .................... |
| V | 5214 hCoV-19/Japan/PG-8391/2020\|EPI_ISL_687490\|2020-07 | .................... | .................... | .................... | .................... | .................... | .................... |
| V | 5215 hCoV-19/Japan/PG-8392/2020\|EPI_ISL_687491\|2020-07 | .................... | .................... | .................... | .................... | .................... | .................... |
| V | 5216 hCoV-19/Japan/PG-8393/2020\|EPI_ISL_687492\|2020-07 | .................... | .................... | .................... | .................... | .................... | .................... |
| V | 5217 hCoV-19/Japan/PG-8394/2020\|EPI_ISL_687493\|2020-07 | .................... | .................... | .................... | .................... | .................... | .................... |
| V | 5218 hCoV-19/Japan/PG-8395/2020\|EPI_ISL_687494\|2020-08 | .................... | .................... | .................... | .................... | .................... | .................... |
| V | 5219 hCoV-19/Japan/PG-8396/2020\|EPI_ISL_687495\|2020-08 | .................... | .................... | .................... | .................... | .................... | .................... |
| V | 5220 hCoV-19/Japan/PG-8397/2020\|EPI_ISL_687496\|2020-08 | .................... | .................... | .................... | .................... | .................... | .................... |
| V | 5221 hCoV-19/Japan/PG-8398/2020\|EPI_ISL_687497\|2020-08 | .................... | .................... | .................... | .................... | .................... | .................... |
| V | 5222 hCoV-19/Japan/PG-8399/2020\|EPI_ISL_687498\|2020-08 | .................... | .................... | .................... | .................... | .................... | .................... |
| V | 5223 hCoV-19/Japan/PG-8400/2020\|EPI_ISL_687499\|2020-08 | .................... | .................... | .................... | .................... | .................... | .................... |
| V | 5224 hCoV-19/Japan/PG-8401/2020\|EPI_ISL_687500\|2020-08 | .................... | .................... | .................... | .................... | .................... | .................... |
| V | 5225 hCoV-19/Japan/PG-8402/2020\|EPI_ISL_687501\|2020-08 | .................... | .................... | .................... | .................... | .................... | .................... |
| V | 5226 hCoV-19/Japan/PG-8403/2020\|EPI_ISL_687502\|2020-08 | .................... | .................... | .................... | .................... | .................... | .................... |
| V | 5227 hCoV-19/Japan/PG-8404/2020\|EPI_ISL_687503\|2020-08 | .................... | .................... | .................... | .................... | .................... | .................... |
| V | 5228 hCoV-19/Japan/PG-8405/2020\|EPI_ISL_687504\|2020-08 | .................... | .................... | .................... | .................... | .................... | .................... |
| V | 5229 hCoV-19/Japan/PG-8406/2020\|EPI_ISL_687505\|2020-08 | .................... | .................... | .................... | .................... | .................... | .................... |
| V | 5230 hCoV-19/Japan/PG-8407/2020\|EPI_ISL_687506\|2020-08 | .................... | .................... | .................... | .................... | .................... | .................... |
| V | 5231 hCoV-19/Japan/PG-8408/2020\|EPI_ISL_687507\|2020-08 | .................... | .................... | .................... | .................... | .................... | .................... |
| V | 5232 hCoV-19/Japan/PG-8409/2020\|EPI_ISL_687508\|2020-08 | .................... | .................... | .................... | .................... | .................... | .................... |
| V | 5233 hCoV-19/Japan/PG-8410/2020\|EPI_ISL_687509\|2020-08 | .................... | .................... | .................... | .................... | .................... | .................... |
| V | 5234 hCoV-19/Japan/PG-5660/2020\|EPI_ISL_688784\|2020-04 | .................... | .................... | .................... | .................... | .................... | .................... |
| V | 5235 hCoV-19/Japan/PG-5661/2020\|EPI_ISL_688785\|2020-04 | .................... | .................... | .................... | .................... | .................... | .................... |
| V | 5236 hCoV-19/Japan/PG-5662/2020\|EPI_ISL_688786\|2020-04 | .................... | .................... | .................... | .................... | .................... | .................... |
| V | 5237 hCoV-19/Japan/PG-5663/2020\|EPI_ISL_688787\|2020-04 | .................... | .................... | .................... | .................... | .................... | .................... |
| V | 5238 hCoV-19/Japan/PG-5664/2020\|EPI_ISL_688788\|2020-04 | .................... | .................... | .................... | .................... | .................... | .................... |
| V | 5239 hCoV-19/Japan/PG-5665/2020\|EPI_ISL_688789\|2020-04 | .................... | .................... | .................... | .................... | .................... | .................... |
| V | 5240 hCoV-19/Japan/PG-5666/2020\|EPI_ISL_688790\|2020-04 | .................... | .................... | .................... | .................... | .................... | .................... |
| V | 5241 hCoV-19/Japan/PG-5667/2020\|EPI_ISL_688791\|2020-04 | .................... | .................... | .................... | .................... | .................... | .................... |
| V | 5242 hCoV-19/Japan/PG-5668/2020\|EPI_ISL_688792\|2020-04 | .................... | .................... | .................... | .................... | .................... | .................... |
| V | 5243 hCoV-19/Japan/PG-5669/2020\|EPI_ISL_688793\|2020-04 | .................... | .................... | .................... | .................... | .................... | .................... |
| V | 5244 hCoV-19/Japan/PG-5670/2020\|EPI_ISL_688794\|2020-04 | .................... | .................... | .................... | .................... | .................... | .................... |
| V | 5245 hCoV-19/Japan/PG-5671/2020\|EPI_ISL_688795\|2020-04 | .................... | .................... | .................... | .................... | .................... | .................... |
| V | 5246 hCoV-19/Japan/PG-5672/2020\|EPI_ISL_688796\|2020-04 | .................... | .................... | .................... | .................... | .................... | .................... |
| V | 5247 hCoV-19/Japan/PG-5673/2020\|EPI_ISL_688797\|2020-04 | .................... | .................... | .................... | .................... | .................... | .................... |
| V | 5248 hCoV-19/Japan/PG-5674/2020\|EPI_ISL_688798\|2020-04 | .................... | .................... | .................... | .................... | .................... | .................... |
| V | 5249 hCoV-19/Japan/PG-5675/2020\|EPI_ISL_688799\|2020-04 | .................... | .................... | .................... | .................... | .................... | .................... |
| V | 5250 hCoV-19/Japan/PG-5676/2020\|EPI_ISL_688800\|2020-04 | .................... | .................... | .................... | .................... | .................... | .................... |
| V | 5251 hCoV-19/Japan/PG-5677/2020\|EPI_ISL_688801\|2020-04 | .................... | .................... | .................... | .................... | .................... | .................... |
| V | 5252 hCoV-19/Japan/PG-5678/2020\|EPI_ISL_688802\|2020-04 | .................... | .................... | .................... | .................... | .................... | .................... |
| V | 5253 hCoV-19/Japan/PG-5679/2020\|EPI_ISL_688803\|2020-04 | .................... | .................... | .................... | .................... | .................... | .................... |
| V | 5254 hCoV-19/Japan/PG-5680/2020\|EPI_ISL_688804\|2020-04 | .................... | .................... | .................... | .................... | .................... | .................... |
| V | 5255 hCoV-19/Japan/PG-5681/2020\|EPI_ISL_688805\|2020-04 | .................... | .................... | .................... | .................... | .................... | .................... |
| V | 5256 hCoV-19/Japan/PG-5682/2020\|EPI_ISL_688806\|2020-04 | .................... | .................... | .................... | .................... | .................... | .................... |
| V | 5257 hCoV-19/Japan/PG-5683/2020\|EPI_ISL_688807\|2020-04 | .................... | .................... | .................... | .................... | .................... | .................... |
| V | 5258 hCoV-19/Japan/PG-5684/2020\|EPI_ISL_688808\|2020-04 | .................... | .A.................. | .................... | .................... | .................... | .................... |
| V | 5259 hCoV-19/Japan/PG-5685/2020\|EPI_ISL_688809\|2020-04 | .................... | .................... | .................... | .................... | .................... | .................... |
| V | 5260 hCoV-19/Japan/PG-5686/2020\|EPI_ISL_688810\|2020-04 | .................... | .A.................. | .................... | .................... | .................... | .................... |
| V | 5261 hCoV-19/Japan/PG-5687/2020\|EPI_ISL_688811\|2020-04 | .................... | .................... | .................... | .................... | .................... | .................... |
| V | 5262 hCoV-19/Japan/PG-5688/2020\|EPI_ISL_688812\|2020-05 | .................... | .................... | .................... | .................... | .................... | .................... |
| V | 5263 hCoV-19/Japan/PG-5689/2020\|EPI_ISL_688813\|2020-05 | .................... | .................... | .................... | .................... | .................... | .................... |
| V | 5264 hCoV-19/Japan/PG-5690/2020\|EPI_ISL_688814\|2020-05 | .................... | .................... | .................... | .................... | .................... | .................... |
| V | 5265 hCoV-19/Japan/PG-5691/2020\|EPI_ISL_688815\|2020-06 | .................... | .................... | .................... | .................... | .................... | .................... |
| V | 5266 hCoV-19/Japan/PG-5692/2020\|EPI_ISL_688816\|2020-06 | .................... | .................... | .................... | .................... | .................... | .................... |
| V | 5267 hCoV-19/Japan/PG-5693/2020\|EPI_ISL_688817\|2020-07 | .................... | .................... | .................... | .................... | .................... | .................... |
| V | 5268 hCoV-19/Japan/PG-5694/2020\|EPI_ISL_688818\|2020-07 | .................... | .................... | .................... | .................... | .................... | .................... |
| V | 5269 hCoV-19/Japan/PG-5695/2020\|EPI_ISL_688819\|2020-07 | .................... | .................... | .................... | .................... | .................... | .................... |
| V | 5270 hCoV-19/Japan/PG-5696/2020\|EPI_ISL_688820\|2020-07 | .................... | .................... | .................... | .................... | .................... | .................... |
| V | 5271 hCoV-19/Japan/PG-5697/2020\|EPI_ISL_688821\|2020-07 | .................... | .................... | .................... | .................... | .................... | .................... |
| V | 5272 hCoV-19/Japan/PG-5698/2020\|EPI_ISL_688822\|2020-07 | .................... | .................... | .................... | .................... | .................... | .................... |
| V | 5273 hCoV-19/Japan/PG-5699/2020\|EPI_ISL_688823\|2020-07 | .................... | .................... | .................... | .................... | .................... | .................... |
| V | 5274 hCoV-19/Japan/PG-5700/2020\|EPI_ISL_688824\|2020-07 | .................... | .................... | .................... | .................... | .................... | .................... |
| V | 5275 hCoV-19/Japan/PG-5701/2020\|EPI_ISL_688825\|2020-07 | .................... | .................... | .................... | .................... | .................... | .................... |
| V | 5276 hCoV-19/Japan/PG-5702/2020\|EPI_ISL_688826\|2020-07 | .................... | .................... | .................... | .................... | .................... | .................... |
| V | 5277 hCoV-19/Japan/PG-5703/2020\|EPI_ISL_688827\|2020-07 | .................... | .................... | .................... | .................... | .................... | .................... |
| V | 5278 hCoV-19/Japan/PG-5705/2020\|EPI_ISL_688828\|2020-08 | .................... | .................... | .................... | .................... | .................... | .................... |
| V | 5279 hCoV-19/Japan/PG-5706/2020\|EPI_ISL_688829\|2020-08 | .................... | .................... | .................... | .................... | .................... | .................... |
| V | 5280 hCoV-19/Japan/PG-5707/2020\|EPI_ISL_688830\|2020-08 | .................... | .................... | .................... | .................... | .................... | .................... |
| V | 5281 hCoV-19/Japan/PG-5708/2020\|EPI_ISL_688831\|2020-08 | .................... | .................... | .................... | .................... | .................... | .................... |
| V | 5282 hCoV-19/Japan/PG-5709/2020\|EPI_ISL_688832\|2020-08 | .................... | .................... | .................... | .................... | .................... | .................... |
| V | 5283 hCoV-19/Japan/PG-5710/2020\|EPI_ISL_688833\|2020-08 | .................... | .................... | .................... | .................... | .................... | .................... |
| V | 5284 hCoV-19/Japan/PG-5711/2020\|EPI_ISL_688834\|2020-08 | .................... | .................... | .................... | .................... | .................... | .................... |
| V | 5285 hCoV-19/Japan/PG-5712/2020\|EPI_ISL_688835\|2020-08 | .................... | .................... | .................... | .................... | .................... | .................... |
| V | 5286 hCoV-19/Japan/PG-5713/2020\|EPI_ISL_688836\|2020-08 | .................... | .................... | .................... | .................... | .................... | .................... |
| V | 5287 hCoV-19/Japan/PG-5714/2020\|EPI_ISL_688837\|2020-08 | .................... | .................... | .................... | .................... | .................... | .................... |
| V | 5288 hCoV-19/Japan/PG-5715/2020\|EPI_ISL_688838\|2020-08 | .................... | .................... | .................... | .................... | .................... | .................... |
| V | 5289 hCoV-19/Japan/PG-5717/2020\|EPI_ISL_688839\|2020-07 | .................... | .................... | .................... | .................... | .................... | .................... |
| V | 5290 hCoV-19/Japan/PG-3758/2020\|EPI_ISL_692011\|2020-04 | .................... | .................... | .................... | .................... | .................... | .................... |

| | | | | | | | |
|---|---|---|---|---|---|---|---|
| V | 5291 hCoV-19/Japan/PG-3759/2020\|EPI_ISL_692012\|2020-04 | ···················· | ···················· | ···················· | ···················· | ···················· | ···················· |
| V | 5292 hCoV-19/Japan/PG-3760/2020\|EPI_ISL_692013\|2020-04 | ···················· | ···················· | ···················· | ···················· | ···················· | ···················· |
| V | 5293 hCoV-19/Japan/PG-3761/2020\|EPI_ISL_692014\|2020-04 | ···················· | ···················· | ···················· | ···················· | ···················· | ···················· |
| V | 5294 hCoV-19/Japan/PG-3762/2020\|EPI_ISL_692015\|2020-04 | ···················· | ···················· | ···················· | ···················· | ···················· | ···················· |
| V | 5295 hCoV-19/Japan/PG-3763/2020\|EPI_ISL_692016\|2020-04 | ···················· | ···················· | ···················· | ···················· | ···················· | ···················· |
| V | 5296 hCoV-19/Japan/PG-3764/2020\|EPI_ISL_692017\|2020-04 | ···················· | ···················· | ···················· | ···················· | ···················· | ···················· |
| V | 5297 hCoV-19/Japan/PG-3765/2020\|EPI_ISL_692018\|2020-04 | ···················· | ···················· | ···················· | ···················· | ···················· | ···················· |
| V | 5298 hCoV-19/Japan/PG-3766/2020\|EPI_ISL_692019\|2020-04 | ···················· | ···················· | ···················· | ···················· | ···················· | ···················· |
| V | 5299 hCoV-19/Japan/PG-3767/2020\|EPI_ISL_692020\|2020-04 | ···················· | ···················· | ···················· | ···················· | ···················· | ···················· |
| V | 5300 hCoV-19/Japan/PG-3768/2020\|EPI_ISL_692021\|2020-04 | ···················· | ···················· | ···················· | ···················· | ···················· | ···················· |
| V | 5301 hCoV-19/Japan/PG-3769/2020\|EPI_ISL_692022\|2020-04 | ···················· | ···················· | ···················· | ···················· | ···················· | ···················· |
| V | 5302 hCoV-19/Japan/PG-3770/2020\|EPI_ISL_692023\|2020-04 | ···················· | ···················· | ···················· | ···················· | ···················· | ···················· |
| V | 5303 hCoV-19/Japan/PG-3771/2020\|EPI_ISL_692024\|2020-04 | ···················· | ···················· | ···················· | ···················· | ···················· | ···················· |
| V | 5304 hCoV-19/Japan/PG-3772/2020\|EPI_ISL_692025\|2020-04 | ···················· | ···················· | ···················· | ···················· | ···················· | ···················· |
| V | 5305 hCoV-19/Japan/PG-3773/2020\|EPI_ISL_692026\|2020-04 | ···················· | ···················· | ···················· | ···················· | ···················· | ···················· |
| V | 5306 hCoV-19/Japan/PG-3775/2020\|EPI_ISL_692027\|2020-04 | ···················· | ···················· | ···················· | ···················· | ···················· | ···················· |
| V | 5307 hCoV-19/Japan/PG-3776/2020\|EPI_ISL_692028\|2020-04 | ···················· | ···················· | ···················· | ···················· | ···················· | ···················· |
| V | 5308 hCoV-19/Japan/PG-3777/2020\|EPI_ISL_692029\|2020-04 | ···················· | ···················· | ···················· | ···················· | ···················· | ···················· |
| V | 5309 hCoV-19/Japan/PG-3778/2020\|EPI_ISL_692030\|2020-04 | ···················· | ···················· | ···················· | ···················· | ···················· | ···················· |
| V | 5310 hCoV-19/Japan/PG-3779/2020\|EPI_ISL_692031\|2020-04 | ···················· | ···················· | ···················· | ···················· | ···················· | ···················· |
| V | 5311 hCoV-19/Japan/PG-3780/2020\|EPI_ISL_692032\|2020-04 | ···················· | ···················· | ···················· | ···················· | ···················· | ···················· |
| V | 5312 hCoV-19/Japan/PG-3782/2020\|EPI_ISL_692033\|2020-04 | ···················· | ···················· | ···················· | ···················· | ···················· | ···················· |
| V | 5313 hCoV-19/Japan/PG-3783/2020\|EPI_ISL_692034\|2020-04 | ···················· | ···················· | ···················· | ···················· | ···················· | ···················· |
| V | 5314 hCoV-19/Japan/PG-3784/2020\|EPI_ISL_692035\|2020-04 | ···················· | ···················· | ···················· | ···················· | ···················· | ···················· |
| V | 5315 hCoV-19/Japan/PG-3785/2020\|EPI_ISL_692036\|2020-04 | ···················· | ···················· | ···················· | ···················· | ···················· | ···················· |
| V | 5316 hCoV-19/Japan/PG-3786/2020\|EPI_ISL_692037\|2020-04 | ···················· | ···················· | ···················· | ···················· | ···················· | ···················· |
| V | 5317 hCoV-19/Japan/PG-3787/2020\|EPI_ISL_692038\|2020-04 | ···················· | ···················· | ···················· | ···················· | ···················· | ···················· |
| V | 5318 hCoV-19/Japan/PG-3788/2020\|EPI_ISL_692039\|2020-04 | ···················· | ···················· | ···················· | ···················· | ···················· | ···················· |
| V | 5319 hCoV-19/Japan/PG-3789/2020\|EPI_ISL_692040\|2020-04 | ···················· | ···················· | ···················· | ···················· | ···················· | ···················· |
| V | 5320 hCoV-19/Japan/PG-3790/2020\|EPI_ISL_692041\|2020-04 | ···················· | ···················· | ···················· | ···················· | ···················· | ···················· |
| V | 5321 hCoV-19/Japan/PG-3791/2020\|EPI_ISL_692042\|2020-04 | ···················· | ···················· | ···················· | ···················· | ···················· | ···················· |
| V | 5322 hCoV-19/Japan/PG-3792/2020\|EPI_ISL_692043\|2020-04 | ···················· | ···················· | ···················· | ···················· | ···················· | ···················· |
| V | 5323 hCoV-19/Japan/PG-3793/2020\|EPI_ISL_692044\|2020-05 | ···················· | ···················· | ···················· | ···················· | ···················· | ···················· |
| V | 5324 hCoV-19/Japan/PG-3794/2020\|EPI_ISL_692045\|2020-05 | ···················· | ···················· | ···················· | ···················· | ···················· | ···················· |
| V | 5325 hCoV-19/Japan/PG-3795/2020\|EPI_ISL_692046\|2020-05 | ···················· | ···················· | ···················· | ···················· | ···················· | ···················· |
| V | 5326 hCoV-19/Japan/PG-3796/2020\|EPI_ISL_692047\|2020-07 | ···················· | ···················· | ···················· | ···················· | ···················· | ···················· |
| V | 5327 hCoV-19/Japan/PG-3797/2020\|EPI_ISL_692048\|2020-07 | ···················· | ·A·················· | ···················· | ···················· | ···················· | ···················· |
| V | 5328 hCoV-19/Japan/PG-3798/2020\|EPI_ISL_692049\|2020-07 | ···················· | ·A·················· | ···················· | ···················· | ···················· | ···················· |
| V | 5329 hCoV-19/Japan/PG-3799/2020\|EPI_ISL_692050\|2020-07 | ···················· | ·A·················· | ···················· | ···················· | ···················· | ···················· |
| V | 5330 hCoV-19/Japan/PG-3800/2020\|EPI_ISL_692051\|2020-07 | ···················· | ···················· | ···················· | ···················· | ···················· | ···················· |
| V | 5331 hCoV-19/Japan/PG-3801/2020\|EPI_ISL_692052\|2020-07 | ···················· | ···················· | ···················· | ···················· | ···················· | ···················· |
| V | 5332 hCoV-19/Japan/PG-3802/2020\|EPI_ISL_692053\|2020-07 | ···················· | ···················· | ···················· | ···················· | ···················· | ···················· |
| V | 5333 hCoV-19/Japan/PG-3803/2020\|EPI_ISL_692054\|2020-07 | ···················· | ···················· | ···················· | ···················· | ···················· | ···················· |
| V | 5334 hCoV-19/Japan/PG-3804/2020\|EPI_ISL_692055\|2020-07 | ···················· | ···················· | ···················· | ···················· | ···················· | ···················· |
| V | 5346 hCoV-19/Thailand/Bangkok-CONI-0274/2020\|EPI_ISL_69 | ···················· | ···················· | ···················· | ···················· | ···················· | ···················· |
| V | 5352 hCoV-19/Thailand/Bangkok-CONI-0280/2020\|EPI_ISL_69 | ···················· | ···················· | ···················· | ···················· | ···················· | ···················· |
| V | 5356 hCoV-19/Thailand/Bangkok-CONI-0290/2020\|EPI_ISL_69 | ···················· | ···················· | ···················· | ···················· | ···················· | ···················· |
| V | 5359 hCoV-19/Thailand/Bangkok-CONI-0294/2020\|EPI_ISL_69 | ···················· | ···················· | ···················· | ···················· | ···················· | ···················· |
| V | 5363 hCoV-19/Thailand/Bangkok-CONI-0299/2020\|EPI_ISL_69 | ···················· | ···················· | ···················· | ···················· | ···················· | ···················· |
| V | 5377 hCoV-19/Japan/PG-2495/2020\|EPI_ISL_686100\|2020-04 | ···················· | ···················· | ···················· | ···················· | ···················· | ···················· |
| V | 5378 hCoV-19/Japan/PG-2496/2020\|EPI_ISL_686101\|2020-04 | ···················· | ···················· | ···················· | ···················· | ···················· | ···················· |
| V | 5379 hCoV-19/Japan/PG-2497/2020\|EPI_ISL_686102\|2020-04 | ···················· | ···················· | ···················· | ···················· | ···················· | ···················· |
| V | 5380 hCoV-19/Japan/PG-2498/2020\|EPI_ISL_686103\|2020-04 | ···················· | ···················· | ···················· | ···················· | ···················· | ···················· |
| V | 5381 hCoV-19/Japan/PG-2499/2020\|EPI_ISL_686104\|2020-04 | ···················· | ···················· | ···················· | ···················· | ···················· | ···················· |
| V | 5382 hCoV-19/Japan/PG-2500/2020\|EPI_ISL_686105\|2020-04 | ···················· | ···················· | ···················· | ···················· | ···················· | ···················· |
| V | 5383 hCoV-19/Japan/PG-2501/2020\|EPI_ISL_686106\|2020-04 | ···················· | ···················· | ···················· | ···················· | ···················· | ···················· |
| V | 5384 hCoV-19/Japan/PG-2502/2020\|EPI_ISL_686107\|2020-04 | ···················· | ···················· | ···················· | ···················· | ···················· | ···················· |
| V | 5385 hCoV-19/Japan/PG-8513/2020\|EPI_ISL_686109\|2020-07 | ···················· | ···················· | ···················· | ···················· | ···················· | ···················· |
| V | 5386 hCoV-19/Japan/PG-8514/2020\|EPI_ISL_686110\|2020-07 | ···················· | ···················· | ···················· | ···················· | ···················· | ···················· |
| V | 5387 hCoV-19/Japan/PG-8515/2020\|EPI_ISL_686111\|2020-07 | ···················· | ···················· | ···················· | ···················· | ···················· | ···················· |
| V | 5388 hCoV-19/Japan/PG-8516/2020\|EPI_ISL_686112\|2020-07 | ···················· | ···················· | ···················· | ···················· | ···················· | ···················· |
| V | 5389 hCoV-19/Japan/PG-8517/2020\|EPI_ISL_686113\|2020-07 | ···················· | ···················· | ···················· | ···················· | ···················· | ···················· |
| V | 5390 hCoV-19/Japan/PG-8518/2020\|EPI_ISL_686114\|2020-07 | ···················· | ···················· | ···················· | ···················· | ···················· | ···················· |
| V | 5391 hCoV-19/Japan/PG-8519/2020\|EPI_ISL_686115\|2020-07 | ···················· | ···················· | ···················· | ···················· | ···················· | ···················· |
| V | 5392 hCoV-19/Japan/PG-8520/2020\|EPI_ISL_686116\|2020-07 | ···················· | ···················· | ···················· | ···················· | ···················· | ···················· |
| V | 5393 hCoV-19/Japan/PG-8521/2020\|EPI_ISL_686117\|2020-07 | ···················· | ···················· | ···················· | ···················· | ···················· | ···················· |
| V | 5394 hCoV-19/Japan/PG-8522/2020\|EPI_ISL_686118\|2020-07 | ···················· | ···················· | ···················· | ···················· | ···················· | ···················· |
| V | 5395 hCoV-19/Japan/PG-8523/2020\|EPI_ISL_686119\|2020-07 | ···················· | ···················· | ···················· | ···················· | ···················· | ···················· |
| V | 5396 hCoV-19/Japan/PG-8524/2020\|EPI_ISL_686120\|2020-07 | ···················· | ···················· | ···················· | ···················· | ···················· | ···················· |
| V | 5397 hCoV-19/Japan/PG-8525/2020\|EPI_ISL_686121\|2020-07 | ···················· | ···················· | ···················· | ···················· | ···················· | ···················· |
| V | 5398 hCoV-19/Japan/PG-8526/2020\|EPI_ISL_686122\|2020-07 | ···················· | ···················· | ···················· | ···················· | ···················· | ···················· |
| V | 5399 hCoV-19/Japan/PG-8584/2020\|EPI_ISL_686180\|2020-08 | ···················· | ···················· | ···················· | ···················· | ···················· | ···················· |
| V | 5400 hCoV-19/Japan/PG-8585/2020\|EPI_ISL_686181\|2020-08 | ···················· | ···················· | ···················· | ···················· | ···················· | ···················· |
| V | 5401 hCoV-19/Japan/PG-8586/2020\|EPI_ISL_686182\|2020-08 | ···················· | ···················· | ···················· | ···················· | ···················· | ···················· |
| V | 5402 hCoV-19/Japan/PG-8587/2020\|EPI_ISL_686183\|2020-08 | ···················· | ···················· | ···················· | ···················· | ···················· | ···················· |
| V | 5403 hCoV-19/Japan/PG-8588/2020\|EPI_ISL_686184\|2020-08 | ···················· | ···················· | ···················· | ···················· | ···················· | ···················· |
| V | 5404 hCoV-19/Japan/PG-8589/2020\|EPI_ISL_686185\|2020-08 | ···················· | ···················· | ···················· | ···················· | ···················· | ···················· |
| V | 5405 hCoV-19/Japan/PG-8590/2020\|EPI_ISL_686186\|2020-08 | ···················· | ···················· | ···················· | ···················· | ···················· | ···················· |
| V | 5406 hCoV-19/Japan/PG-8591/2020\|EPI_ISL_686187\|2020-08 | ···················· | ···················· | ···················· | ···················· | ···················· | ···················· |
| V | 5407 hCoV-19/Japan/PG-8592/2020\|EPI_ISL_686188\|2020-08 | ···················· | ···················· | ···················· | ···················· | ···················· | ···················· |

| | | | | | | | | |
|---|---|---|---|---|---|---|---|---|
| V | 5408 | hCoV-19/Japan/PG-8593/2020\|EPI_ISL_686189\|2020-08 | .................... | .................... | .................... | .................... | .................... | .................... |
| V | 5409 | hCoV-19/Japan/PG-8595/2020\|EPI_ISL_686191\|2020-08 | .................... | .................... | .................... | .................... | .................... | .................... |
| V | 5410 | hCoV-19/Japan/PG-8596/2020\|EPI_ISL_686192\|2020-08 | .................... | .................... | .................... | .................... | .................... | .................... |
| V | 5411 | hCoV-19/Japan/PG-8597/2020\|EPI_ISL_686193\|2020-08 | .................... | .................... | .................... | .................... | .................... | .................... |
| V | 5412 | hCoV-19/Japan/PG-8598/2020\|EPI_ISL_686194\|2020-08 | .................... | .................... | .................... | .................... | .................... | .................... |
| V | 5413 | hCoV-19/Japan/PG-8599/2020\|EPI_ISL_686195\|2020-08 | .................... | .................... | .................... | .................... | .................... | .................... |
| V | 5414 | hCoV-19/Japan/PG-8600/2020\|EPI_ISL_686196\|2020-08 | .................... | .................... | .................... | .................... | .................... | .................... |
| V | 5415 | hCoV-19/Japan/PG-8601/2020\|EPI_ISL_686197\|2020-08 | .................... | .................... | .................... | .................... | .................... | .................... |
| V | 5416 | hCoV-19/Japan/PG-8602/2020\|EPI_ISL_686198\|2020-08 | .................... | .................... | .................... | .................... | .................... | .................... |
| V | 5417 | hCoV-19/Japan/PG-8603/2020\|EPI_ISL_686199\|2020-08 | .................... | .................... | .................... | .................... | .................... | .................... |
| V | 5418 | hCoV-19/Japan/PG-8301/2020\|EPI_ISL_687400\|2020-08 | .................... | .................... | .................... | .................... | .................... | .................... |
| V | 5419 | hCoV-19/Japan/PG-8302/2020\|EPI_ISL_687401\|2020-08 | .................... | .................... | .................... | .................... | .................... | .................... |
| V | 5420 | hCoV-19/Japan/PG-8303/2020\|EPI_ISL_687402\|2020-08 | .................... | .................... | .................... | .................... | .................... | .................... |
| V | 5421 | hCoV-19/Japan/PG-8304/2020\|EPI_ISL_687403\|2020-08 | .................... | .................... | .................... | .................... | .................... | .................... |
| V | 5422 | hCoV-19/Japan/PG-8305/2020\|EPI_ISL_687404\|2020-08 | .................... | .................... | .................... | .................... | .................... | .................... |
| V | 5423 | hCoV-19/Japan/PG-8306/2020\|EPI_ISL_687405\|2020-08 | .................... | .................... | .................... | .................... | .................... | .................... |
| V | 5424 | hCoV-19/Japan/PG-8307/2020\|EPI_ISL_687406\|2020-08 | .................... | .................... | .................... | .................... | .................... | .................... |
| V | 5425 | hCoV-19/Japan/PG-8308/2020\|EPI_ISL_687407\|2020-08 | .................... | .................... | .................... | .................... | .................... | .................... |
| V | 5426 | hCoV-19/Japan/PG-8309/2020\|EPI_ISL_687408\|2020-08 | .................... | .................... | .................... | .................... | .................... | .................... |
| V | 5427 | hCoV-19/Japan/PG-8310/2020\|EPI_ISL_687409\|2020-08 | .................... | .................... | .................... | .................... | .................... | .................... |
| V | 5428 | hCoV-19/Japan/PG-8311/2020\|EPI_ISL_687410\|2020-08 | .................... | .................... | .................... | .................... | .................... | .................... |
| V | 5429 | hCoV-19/Japan/PG-8312/2020\|EPI_ISL_687411\|2020-08 | .................... | .................... | .................... | .................... | .................... | .................... |
| V | 5430 | hCoV-19/Japan/PG-8313/2020\|EPI_ISL_687412\|2020-08 | .................... | .................... | .................... | .................... | .................... | .................... |
| V | 5431 | hCoV-19/Japan/PG-8314/2020\|EPI_ISL_687413\|2020-08 | .................... | .................... | .................... | .................... | .................... | .................... |
| V | 5432 | hCoV-19/Japan/PG-8315/2020\|EPI_ISL_687414\|2020-08 | .................... | .................... | .................... | .................... | .................... | .................... |
| V | 5433 | hCoV-19/Japan/PG-8316/2020\|EPI_ISL_687415\|2020-08 | .................... | .................... | .................... | .................... | .................... | .................... |
| V | 5434 | hCoV-19/Japan/PG-8317/2020\|EPI_ISL_687416\|2020-08 | .................... | .................... | .................... | .................... | .................... | .................... |
| V | 5435 | hCoV-19/Japan/PG-8318/2020\|EPI_ISL_687417\|2020-08 | .................... | .................... | .................... | .................... | .................... | .................... |
| V | 5436 | hCoV-19/Japan/PG-8319/2020\|EPI_ISL_687418\|2020-08 | .................... | .................... | .................... | .................... | .................... | .................... |
| V | 5437 | hCoV-19/Japan/PG-8320/2020\|EPI_ISL_687419\|2020-08 | .................... | .................... | .................... | .................... | .................... | .................... |
| V | 5438 | hCoV-19/Japan/PG-8321/2020\|EPI_ISL_687420\|2020-08 | .................... | .................... | .................... | .................... | .................... | .................... |
| V | 5439 | hCoV-19/Japan/PG-8322/2020\|EPI_ISL_687421\|2020-08 | .................... | .................... | .................... | .................... | .................... | .................... |
| V | 5440 | hCoV-19/Japan/PG-8323/2020\|EPI_ISL_687422\|2020-08 | .................... | .................... | .................... | .................... | .................... | .................... |
| V | 5441 | hCoV-19/Japan/PG-8324/2020\|EPI_ISL_687423\|2020-08 | .................... | .................... | .................... | .................... | .................... | .................... |
| V | 5442 | hCoV-19/Japan/PG-8325/2020\|EPI_ISL_687424\|2020-08 | .................... | .................... | .................... | .................... | .................... | .................... |
| V | 5443 | hCoV-19/Japan/PG-8326/2020\|EPI_ISL_687425\|2020-08 | .................... | .................... | .................... | .................... | .................... | .................... |
| V | 5444 | hCoV-19/Japan/PG-8327/2020\|EPI_ISL_687426\|2020-08 | .................... | .................... | .................... | .................... | .................... | .................... |
| V | 5445 | hCoV-19/Japan/PG-8328/2020\|EPI_ISL_687427\|2020-08 | .................... | .................... | .................... | .................... | .................... | .................... |
| V | 5446 | hCoV-19/Japan/PG-8329/2020\|EPI_ISL_687428\|2020-08 | .................... | .................... | .................... | .................... | .................... | .................... |
| V | 5447 | hCoV-19/Japan/PG-8330/2020\|EPI_ISL_687429\|2020-08 | .................... | .................... | .................... | .................... | .................... | .................... |
| V | 5448 | hCoV-19/Japan/PG-8331/2020\|EPI_ISL_687430\|2020-08 | .................... | .................... | .................... | .................... | .................... | .................... |
| V | 5449 | hCoV-19/Japan/PG-8332/2020\|EPI_ISL_687431\|2020-08 | .................... | .................... | .................... | .................... | .................... | .................... |
| V | 5450 | hCoV-19/Japan/PG-8333/2020\|EPI_ISL_687432\|2020-08 | .................... | .................... | .................... | .................... | .................... | .................... |
| V | 5451 | hCoV-19/Japan/PG-8334/2020\|EPI_ISL_687433\|2020-08 | .................... | .................... | .................... | .................... | .................... | .................... |
| V | 5452 | hCoV-19/Japan/PG-8335/2020\|EPI_ISL_687434\|2020-08 | .................... | .................... | .................... | .................... | .................... | .................... |
| V | 5453 | hCoV-19/Japan/PG-8336/2020\|EPI_ISL_687435\|2020-08 | .................... | .................... | .................... | .................... | .................... | .................... |
| V | 5454 | hCoV-19/Japan/PG-8337/2020\|EPI_ISL_687436\|2020-08 | .................... | .................... | .................... | .................... | .................... | .................... |
| V | 5455 | hCoV-19/Japan/PG-8338/2020\|EPI_ISL_687437\|2020-08 | .................... | .................... | .................... | .................... | .................... | .................... |
| V | 5456 | hCoV-19/Japan/PG-8339/2020\|EPI_ISL_687438\|2020-08 | .................... | .................... | .................... | .................... | .................... | .................... |
| V | 5457 | hCoV-19/Japan/PG-8340/2020\|EPI_ISL_687439\|2020-08 | .................... | .................... | .................... | .................... | .................... | .................... |
| V | 5458 | hCoV-19/Japan/PG-8341/2020\|EPI_ISL_687440\|2020-08 | .................... | .................... | .................... | .................... | .................... | .................... |
| V | 5459 | hCoV-19/Japan/PG-8342/2020\|EPI_ISL_687441\|2020-08 | .................... | .................... | .................... | .................... | .................... | .................... |
| V | 5460 | hCoV-19/Japan/PG-8343/2020\|EPI_ISL_687442\|2020-08 | .................... | .................... | .................... | .................... | .................... | .................... |
| V | 5461 | hCoV-19/Japan/PG-8344/2020\|EPI_ISL_687443\|2020-08 | .................... | .................... | .................... | .................... | .................... | .................... |
| V | 5462 | hCoV-19/Japan/PG-8345/2020\|EPI_ISL_687444\|2020-08 | .................... | .................... | .................... | .................... | .................... | .................... |
| V | 5463 | hCoV-19/Japan/PG-8346/2020\|EPI_ISL_687445\|2020-08 | .................... | .................... | .................... | .................... | .................... | .................... |
| V | 5464 | hCoV-19/Japan/PG-8347/2020\|EPI_ISL_687446\|2020-08 | .................... | .................... | .................... | .................... | .................... | .................... |
| V | 5465 | hCoV-19/Japan/PG-8348/2020\|EPI_ISL_687447\|2020-08 | .................... | .................... | .................... | .................... | .................... | .................... |
| V | 5466 | hCoV-19/Japan/PG-8349/2020\|EPI_ISL_687448\|2020-08 | .................... | .................... | .................... | .................... | .................... | .................... |
| V | 5467 | hCoV-19/Japan/PG-8350/2020\|EPI_ISL_687449\|2020-08 | .................... | .................... | .................... | .................... | .................... | .................... |
| V | 5468 | hCoV-19/Japan/PG-8351/2020\|EPI_ISL_687450\|2020-08 | .................... | .................... | .................... | .................... | .................... | .................... |
| V | 5469 | hCoV-19/Japan/PG-8352/2020\|EPI_ISL_687451\|2020-08 | .................... | .................... | .................... | .................... | .................... | .................... |
| V | 5470 | hCoV-19/Japan/PG-8353/2020\|EPI_ISL_687452\|2020-08 | .................... | .................... | .................... | .................... | .................... | .................... |
| V | 5471 | hCoV-19/Japan/PG-8354/2020\|EPI_ISL_687453\|2020-08 | .................... | .................... | .................... | .................... | .................... | .................... |
| V | 5472 | hCoV-19/Japan/PG-5575/2020\|EPI_ISL_688700\|2020-08 | .................... | .................... | .................... | .................... | .................... | .................... |
| V | 5473 | hCoV-19/Japan/PG-5576/2020\|EPI_ISL_688701\|2020-08 | .................... | .................... | .................... | .................... | .................... | .................... |
| V | 5474 | hCoV-19/Japan/PG-5577/2020\|EPI_ISL_688702\|2020-08 | .................... | .................... | .................... | .................... | .................... | .................... |
| V | 5475 | hCoV-19/Japan/PG-5578/2020\|EPI_ISL_688703\|2020-08 | .................... | .................... | .................... | .................... | .................... | .................... |
| V | 5476 | hCoV-19/Japan/PG-5579/2020\|EPI_ISL_688704\|2020-08 | .................... | .................... | .................... | .................... | .................... | .................... |
| V | 5477 | hCoV-19/Japan/PG-5580/2020\|EPI_ISL_688705\|2020-08 | .................... | .................... | .................... | .................... | .................... | .................... |
| V | 5478 | hCoV-19/Japan/PG-5581/2020\|EPI_ISL_688706\|2020-08 | .................... | .................... | .................... | .................... | .................... | .................... |
| V | 5479 | hCoV-19/Japan/PG-5582/2020\|EPI_ISL_688707\|2020-08 | .................... | .................... | .................... | .................... | .................... | .................... |
| V | 5480 | hCoV-19/Japan/PG-5583/2020\|EPI_ISL_688708\|2020-08 | .................... | .................... | .................... | .................... | .................... | .................... |
| V | 5481 | hCoV-19/Japan/PG-5584/2020\|EPI_ISL_688709\|2020-08 | .................... | .................... | .................... | .................... | .................... | .................... |
| V | 5482 | hCoV-19/Japan/PG-5585/2020\|EPI_ISL_688710\|2020-08 | .................... | .................... | .................... | .................... | .................... | .................... |
| V | 5483 | hCoV-19/Japan/PG-5586/2020\|EPI_ISL_688711\|2020-08 | .................... | .................... | .................... | .................... | .................... | .................... |
| V | 5484 | hCoV-19/Japan/PG-5587/2020\|EPI_ISL_688712\|2020-08 | .................... | .................... | .................... | .................... | .................... | .................... |
| V | 5485 | hCoV-19/Japan/PG-5588/2020\|EPI_ISL_688713\|2020-08 | .................... | .................... | .................... | .................... | .................... | .................... |
| V | 5486 | hCoV-19/Japan/PG-5589/2020\|EPI_ISL_688714\|2020-08 | .................... | .................... | .................... | .................... | .................... | .................... |
| V | 5487 | hCoV-19/Japan/PG-5590/2020\|EPI_ISL_688715\|2020-08 | .................... | .................... | .................... | .................... | .................... | .................... |

| | | | | | | | | |
|---|---|---|---|---|---|---|---|---|
| V | 5488 | hCoV-19/Japan/PG-5591/2020\|EPI_ISL_688716\|2020-08 | ···················· | ···················· | ····················· | ···················· | ···················· | ···················· |
| V | 5489 | hCoV-19/Japan/PG-5592/2020\|EPI_ISL_688717\|2020-08 | ···················· | ···················· | ····················· | ···················· | ···················· | ···················· |
| V | 5490 | hCoV-19/Japan/PG-5593/2020\|EPI_ISL_688718\|2020-08 | ···················· | ···················· | ····················· | ···················· | ···················· | ···················· |
| V | 5491 | hCoV-19/Japan/PG-5594/2020\|EPI_ISL_688719\|2020-08 | ···················· | ···················· | ····················· | ···················· | ···················· | ···················· |
| V | 5492 | hCoV-19/Japan/PG-5595/2020\|EPI_ISL_688720\|2020-08 | ···················· | ···················· | ····················· | ···················· | ···················· | ···················· |
| V | 5493 | hCoV-19/Japan/PG-5596/2020\|EPI_ISL_688721\|2020-08 | ···················· | ···················· | ····················· | ···················· | ···················· | ···················· |
| V | 5494 | hCoV-19/Japan/PG-5597/2020\|EPI_ISL_688722\|2020-08 | ···················· | ···················· | ····················· | ···················· | ···················· | ···················· |
| V | 5495 | hCoV-19/Japan/PG-5598/2020\|EPI_ISL_688723\|2020-08 | ···················· | ···················· | ····················· | ···················· | ···················· | ···················· |
| V | 5496 | hCoV-19/Japan/PG-5599/2020\|EPI_ISL_688724\|2020-08 | ···················· | ···················· | ····················· | ···················· | ···················· | ···················· |
| V | 5497 | hCoV-19/Japan/PG-5600/2020\|EPI_ISL_688725\|2020-08 | ···················· | ···················· | ····················· | ···················· | ···················· | ···················· |
| V | 5498 | hCoV-19/Japan/PG-5601/2020\|EPI_ISL_688726\|2020-08 | ···················· | ···················· | ····················· | ···················· | ···················· | ···················· |
| V | 5499 | hCoV-19/Japan/PG-5602/2020\|EPI_ISL_688727\|2020-08 | ···················· | ···················· | ····················· | ···················· | ···················· | ···················· |
| V | 5500 | hCoV-19/Japan/PG-5603/2020\|EPI_ISL_688728\|2020-08 | ···················· | ···················· | ····················· | ···················· | ···················· | ···················· |
| V | 5501 | hCoV-19/Japan/PG-5604/2020\|EPI_ISL_688729\|2020-08 | ···················· | ···················· | ····················· | ···················· | ···················· | ···················· |
| V | 5502 | hCoV-19/Japan/PG-5605/2020\|EPI_ISL_688730\|2020-08 | ···················· | ···················· | ····················· | ···················· | ···················· | ···················· |
| V | 5503 | hCoV-19/Japan/PG-5606/2020\|EPI_ISL_688731\|2020-08 | ···················· | ···················· | ····················· | ···················· | ···················· | ···················· |
| V | 5504 | hCoV-19/Japan/PG-5607/2020\|EPI_ISL_688732\|2020-08 | ···················· | ···················· | ····················· | ···················· | ···················· | ···················· |
| V | 5505 | hCoV-19/Japan/PG-5608/2020\|EPI_ISL_688733\|2020-08 | ···················· | ···················· | ····················· | ···················· | ···················· | ···················· |
| V | 5506 | hCoV-19/Japan/PG-5609/2020\|EPI_ISL_688734\|2020-08 | ···················· | ···················· | ····················· | ···················· | ···················· | ···················· |
| V | 5507 | hCoV-19/Japan/PG-5610/2020\|EPI_ISL_688735\|2020-08 | ···················· | ···················· | ····················· | ···················· | ···················· | ···················· |
| V | 5508 | hCoV-19/Japan/PG-5611/2020\|EPI_ISL_688736\|2020-08 | ···················· | ···················· | ····················· | ···················· | ···················· | ···················· |
| V | 5509 | hCoV-19/Japan/PG-5612/2020\|EPI_ISL_688737\|2020-08 | ···················· | ···················· | ····················· | ···················· | ···················· | ···················· |
| V | 5510 | hCoV-19/Japan/PG-5613/2020\|EPI_ISL_688738\|2020-07 | ···················· | ···················· | ····················· | ···················· | ···················· | ···················· |
| V | 5511 | hCoV-19/Japan/PG-5614/2020\|EPI_ISL_688739\|2020-07 | ···················· | ···················· | ····················· | ···················· | ···················· | ···················· |
| V | 5512 | hCoV-19/Japan/PG-5615/2020\|EPI_ISL_688740\|2020-07 | ···················· | ···················· | ····················· | ···················· | ···················· | ···················· |
| V | 5513 | hCoV-19/Japan/PG-5616/2020\|EPI_ISL_688741\|2020-08 | ···················· | ···················· | ····················· | ···················· | ···················· | ···················· |
| V | 5514 | hCoV-19/Japan/PG-5617/2020\|EPI_ISL_688742\|2020-08 | ···················· | ···················· | ····················· | ···················· | ···················· | ···················· |
| V | 5515 | hCoV-19/Japan/PG-5618/2020\|EPI_ISL_688743\|2020-08 | ···················· | ···················· | ····················· | ···················· | ···················· | ···················· |
| V | 5516 | hCoV-19/Japan/PG-5619/2020\|EPI_ISL_688744\|2020-08 | ···················· | ···················· | ····················· | ···················· | ···················· | ···················· |
| V | 5517 | hCoV-19/Japan/PG-5620/2020\|EPI_ISL_688745\|2020-08 | ···················· | ···················· | ····················· | ···················· | ···················· | ···················· |
| V | 5518 | hCoV-19/Japan/PG-5621/2020\|EPI_ISL_688746\|2020-09 | ···················· | ···················· | ····················· | ···················· | ···················· | ···················· |
| V | 5519 | hCoV-19/Japan/PG-5622/2020\|EPI_ISL_688747\|2020-08 | ···················· | ···················· | ····················· | ···················· | ···················· | ···················· |
| V | 5520 | hCoV-19/Japan/PG-5623/2020\|EPI_ISL_688748\|2020-08 | ···················· | ···················· | ····················· | ···················· | ···················· | ···················· |
| V | 5521 | hCoV-19/Japan/PG-5624/2020\|EPI_ISL_688749\|2020-08 | ···················· | ···················· | ····················· | ···················· | ···················· | ···················· |
| V | 5522 | hCoV-19/Japan/PG-5625/2020\|EPI_ISL_688750\|2020-08 | ···················· | ···················· | ····················· | ···················· | ···················· | ···················· |
| V | 5523 | hCoV-19/Japan/PG-5626/2020\|EPI_ISL_688751\|2020-08 | ···················· | ···················· | ····················· | ···················· | ···················· | ···················· |
| V | 5524 | hCoV-19/Japan/PG-5627/2020\|EPI_ISL_688752\|2020-08 | ···················· | ···················· | ····················· | ···················· | ···················· | ···················· |
| V | 5525 | hCoV-19/Japan/PG-5628/2020\|EPI_ISL_688753\|2020-08 | ···················· | ···················· | ····················· | ···················· | ···················· | ···················· |
| V | 5526 | hCoV-19/Japan/PG-5629/2020\|EPI_ISL_688754\|2020-08 | ···················· | ···················· | ····················· | ···················· | ···················· | ···················· |
| V | 5527 | hCoV-19/Japan/PG-5630/2020\|EPI_ISL_688755\|2020-08 | ···················· | ···················· | ····················· | ···················· | ···················· | ···················· |
| V | 5528 | hCoV-19/Japan/PG-5631/2020\|EPI_ISL_688756\|2020-08 | ···················· | ···················· | ····················· | ···················· | ···················· | ···················· |
| V | 5529 | hCoV-19/Japan/PG-5632/2020\|EPI_ISL_688757\|2020-03 | ···················· | ···················· | ····················· | ···················· | ···················· | ···················· |
| V | 5530 | hCoV-19/Japan/PG-5633/2020\|EPI_ISL_688758\|2020-03 | ···················· | ···················· | ····················· | ···················· | ···················· | ···················· |
| V | 5531 | hCoV-19/Japan/PG-5634/2020\|EPI_ISL_688759\|2020-03 | ···················· | ···················· | ····················· | ···················· | ···················· | ···················· |
| V | 5532 | hCoV-19/Japan/PG-5635/2020\|EPI_ISL_688760\|2020-03 | ···················· | ···················· | ····················· | ···················· | ···················· | ···················· |
| V | 5533 | hCoV-19/Japan/PG-5637/2020\|EPI_ISL_688761\|2020-03 | ···················· | ···················· | ····················· | ···················· | ···················· | ···················· |
| V | 5534 | hCoV-19/Japan/PG-5638/2020\|EPI_ISL_688762\|2020-04 | ···················· | ···················· | ····················· | ···················· | ···················· | ···················· |
| V | 5535 | hCoV-19/Japan/PG-5639/2020\|EPI_ISL_688763\|2020-04 | ···················· | ···················· | ····················· | ···················· | ···················· | ···················· |
| V | 5536 | hCoV-19/Japan/PG-5640/2020\|EPI_ISL_688764\|2020-04 | ···················· | ···················· | ····················· | ···················· | ···················· | ···················· |
| V | 5537 | hCoV-19/Japan/PG-5641/2020\|EPI_ISL_688765\|2020-04 | ···················· | ···················· | ····················· | ···················· | ···················· | ···················· |
| V | 5538 | hCoV-19/Japan/PG-5642/2020\|EPI_ISL_688766\|2020-04 | ···················· | ···················· | ····················· | ···················· | ···················· | ···················· |
| V | 5539 | hCoV-19/Japan/PG-5643/2020\|EPI_ISL_688767\|2020-04 | ···················· | ···················· | ····················· | ···················· | ···················· | ···················· |
| V | 5540 | hCoV-19/Japan/PG-5644/2020\|EPI_ISL_688768\|2020-04 | ···················· | ···················· | ····················· | ···················· | ···················· | ···················· |
| V | 5541 | hCoV-19/Japan/PG-5645/2020\|EPI_ISL_688769\|2020-04 | ···················· | ···················· | ····················· | ···················· | ···················· | ···················· |
| V | 5542 | hCoV-19/Japan/PG-5646/2020\|EPI_ISL_688770\|2020-04 | ···················· | ···················· | ····················· | ···················· | ···················· | ···················· |
| V | 5543 | hCoV-19/Japan/PG-5647/2020\|EPI_ISL_688771\|2020-04 | ···················· | ···················· | ····················· | ···················· | ···················· | ···················· |
| V | 5544 | hCoV-19/Japan/PG-5648/2020\|EPI_ISL_688772\|2020-04 | ···················· | ···················· | ····················· | ···················· | ···················· | ···················· |
| V | 5545 | hCoV-19/Japan/PG-5649/2020\|EPI_ISL_688773\|2020-04 | ···················· | ···················· | ····················· | ···················· | ···················· | ···················· |
| V | 5546 | hCoV-19/Japan/PG-5650/2020\|EPI_ISL_688774\|2020-04 | ···················· | ···················· | ····················· | ···················· | ···················· | ···················· |
| V | 5547 | hCoV-19/Japan/PG-5651/2020\|EPI_ISL_688775\|2020-04 | ···················· | ···················· | ····················· | ···················· | ···················· | ···················· |
| V | 5548 | hCoV-19/Japan/PG-5652/2020\|EPI_ISL_688776\|2020-04 | ···················· | ···················· | ····················· | ···················· | ···················· | ···················· |
| V | 5549 | hCoV-19/Japan/PG-5653/2020\|EPI_ISL_688777\|2020-04 | ···················· | ···················· | ····················· | ···················· | ···················· | ···················· |
| V | 5550 | hCoV-19/Japan/PG-5654/2020\|EPI_ISL_688778\|2020-04 | ···················· | ···················· | ····················· | ···················· | ···················· | ···················· |
| V | 5551 | hCoV-19/Japan/PG-5655/2020\|EPI_ISL_688779\|2020-04 | ···················· | ···················· | ····················· | ···················· | ···················· | ···················· |
| V | 5552 | hCoV-19/Japan/PG-5656/2020\|EPI_ISL_688780\|2020-04 | ···················· | ···················· | ····················· | ···················· | ···················· | ···················· |
| V | 5553 | hCoV-19/Japan/PG-5657/2020\|EPI_ISL_688781\|2020-04 | ···················· | ···················· | ····················· | ···················· | ···················· | ···················· |
| V | 5554 | hCoV-19/Japan/PG-5658/2020\|EPI_ISL_688782\|2020-04 | ···················· | ···················· | ····················· | ···················· | ···················· | ···················· |
| V | 5555 | hCoV-19/Japan/PG-5659/2020\|EPI_ISL_688783\|2020-04 | ···················· | ···················· | ····················· | ···················· | ···················· | ···················· |
| V | 5556 | hCoV-19/Brazil/SP-571/2020\|EPI_ISL_693200\|2020-04- | ···················· | ···················· | ····················· | ···················· | ···················· | ···················· |
| V | 5557 | hCoV-19/Brazil/SP-572/2020\|EPI_ISL_693201\|2020-04- | ···················· | ···················· | ····················· | ···················· | ···················· | ···················· |
| V | 5558 | hCoV-19/Brazil/SP-573/2020\|EPI_ISL_693202\|2020-04- | ···················· | ···················· | ····················· | ···················· | ···················· | ···················· |
| V | 5559 | hCoV-19/Brazil/SP-575/2020\|EPI_ISL_693203\|2020-04- | ···················· | ···················· | ····················· | ···················· | ···················· | ···················· |
| V | 5560 | hCoV-19/Brazil/SP-352/2020\|EPI_ISL_693204\|2020-06- | ···················· | ···················· | ····················· | ···················· | ···················· | ···················· |
| V | 5561 | hCoV-19/Brazil/SP-577/2020\|EPI_ISL_693205\|2020-06- | ···················· | ···················· | ····················· | ···················· | ···················· | ···················· |
| V | 5562 | hCoV-19/Brazil/SP-361/2020\|EPI_ISL_693206\|2020-06- | ···················· | ···················· | ····················· | ···················· | ···················· | ···················· |
| V | 5563 | hCoV-19/Brazil/SP-366/2020\|EPI_ISL_693207\|2020-06- | ···················· | ···················· | ····················· | ···················· | ···················· | ···················· |
| V | 5564 | hCoV-19/Brazil/SP-400/2020\|EPI_ISL_693208\|2020-07- | ··C················· | ···················· | ····················· | ···················· | ···················· | ···················· |
| V | 5565 | hCoV-19/Brazil/SP-401/2020\|EPI_ISL_693209\|2020-07- | ···················· | ···················· | ····················· | ···················· | ···················· | ···················· |
| V | 5566 | hCoV-19/Brazil/SP-415/2020\|EPI_ISL_693210\|2020-07- | ···················· | ···················· | ····················· | ···················· | ···················· | ···················· |
| V | 5567 | hCoV-19/Brazil/SP-416/2020\|EPI_ISL_693211\|2020-07- | ···················· | ···················· | ····················· | ···················· | ···················· | ···················· |

| | | | | | | | | |
|---|---|---|---|---|---|---|---|---|
| V | 5568 | hCoV-19/Brazil/SP-423/2020\|EPI_ISL_693212\|2020-07- | ···················· | ···················· | ····················· | ···················· | ···················· | ···················· |
| V | 5569 | hCoV-19/Brazil/SP-390/2020\|EPI_ISL_693213\|2020-06- | ···················· | ···················· | ····················· | ···················· | ···················· | ···················· |
| V | 5570 | hCoV-19/Brazil/SP-579/2020\|EPI_ISL_693214\|2020-07- | ···················· | ···················· | ····················· | ···················· | ···················· | ···················· |
| V | 5571 | hCoV-19/Brazil/SP-589/2020\|EPI_ISL_693215\|2020-07- | ···················· | ···················· | ····················· | ···················· | ···················· | ···················· |
| V | 5572 | hCoV-19/Brazil/SP-590/2020\|EPI_ISL_693216\|2020-07- | ···················· | ···················· | ····················· | ···················· | ···················· | ···················· |
| V | 5573 | hCoV-19/Brazil/SP-591/2020\|EPI_ISL_693217\|2020-07- | ···················· | ···················· | ····················· | ···················· | ···················· | ···················· |
| V | 5574 | hCoV-19/Brazil/SP-594/2020\|EPI_ISL_693218\|2020-06- | ···················· | ···················· | ····················· | ···················· | ···················· | ···················· |
| V | 5575 | hCoV-19/Brazil/SP-595/2020\|EPI_ISL_693219\|2020-06- | ···················· | ···················· | ····················· | ···················· | ···················· | ···················· |
| V | 5576 | hCoV-19/Brazil/SP-596/2020\|EPI_ISL_693220\|2020-07- | ···················· | ···················· | ····················· | ···················· | ···················· | ···················· |
| V | 5577 | hCoV-19/Brazil/SP-597/2020\|EPI_ISL_693221\|2020-06- | ···················· | ···················· | ····················· | ···················· | ···················· | ···················· |
| V | 5578 | hCoV-19/Brazil/SP-598/2020\|EPI_ISL_693222\|2020-06- | ···················· | ···················· | ···········T········· | ···················· | ···················· | ···················· |
| V | 5579 | hCoV-19/Brazil/SP-643/2020\|EPI_ISL_693223\|2020-07- | ···················· | ···················· | ····················· | ···················· | ···················· | ···················· |
| V | 5580 | hCoV-19/Brazil/SP-645/2020\|EPI_ISL_693224\|2020-07- | ···················· | ···················· | ····················· | ···················· | ···················· | ···················· |
| V | 5581 | hCoV-19/Brazil/SP-652/2020\|EPI_ISL_693225\|2020-07- | ···················· | ···················· | ····················· | ···················· | ···················· | ···················· |
| V | 5582 | hCoV-19/Brazil/SP-653/2020\|EPI_ISL_693226\|2020-06- | ···················· | ···················· | ····················· | ···················· | ···················· | ···················· |
| V | 5583 | hCoV-19/Brazil/SP-658/2020\|EPI_ISL_693227\|2020-07- | ···················· | ···················· | ····················· | ···················· | ···················· | ···················· |
| V | 5584 | hCoV-19/Brazil/SP-661/2020\|EPI_ISL_693228\|2020-06- | ···················· | ···················· | ····················· | ···················· | ···················· | ···················· |
| V | 5585 | hCoV-19/Brazil/SP-666/2020\|EPI_ISL_693229\|2020-06- | ···················· | ···················· | ····················· | ···················· | ···················· | ···················· |
| V | 5586 | hCoV-19/Brazil/SP-667/2020\|EPI_ISL_693230\|2020-06- | ···················· | ···················· | ····················· | ···················· | ···················· | ···················· |
| V | 5587 | hCoV-19/Brazil/SP-668/2020\|EPI_ISL_693231\|2020-06- | ···················· | ···················· | ····················· | ···················· | ···················· | ···················· |
| V | 5588 | hCoV-19/Brazil/SP-673/2020\|EPI_ISL_693232\|2020-05- | ···················· | ····G··············· | ····················· | ···················· | ···················· | ···················· |
| V | 5589 | hCoV-19/Brazil/SP-678/2020\|EPI_ISL_693233\|2020-06- | ···················· | ···················· | ····················· | ···················· | ···················· | ···················· |
| V | 5590 | hCoV-19/Brazil/SP-688/2020\|EPI_ISL_693234\|2020-06- | ···················· | ···················· | ····················· | ···················· | ···················· | ···················· |
| V | 5591 | hCoV-19/Brazil/SP-689/2020\|EPI_ISL_693235\|2020-06- | ···················· | ···················· | ····················· | ···················· | ···················· | ···················· |
| V | 5592 | hCoV-19/Brazil/SP-692/2020\|EPI_ISL_693236\|2020-07- | ···················· | ···················· | ····················· | ···················· | ···················· | ···················· |
| V | 5593 | hCoV-19/Brazil/SP-694/2020\|EPI_ISL_693237\|2020-07- | ···················· | ···················· | ····················· | ···················· | ···················· | ···················· |
| V | 5594 | hCoV-19/Brazil/SP-696/2020\|EPI_ISL_693238\|2020-06- | ···················· | ···················· | ····················· | ···················· | ···················· | ···················· |
| V | 5595 | hCoV-19/Brazil/SP-697/2020\|EPI_ISL_693239\|2020-06- | ···················· | ···················· | ····················· | ···················· | ···················· | ···················· |
| V | 5596 | hCoV-19/Brazil/SP-699/2020\|EPI_ISL_693241\|2020-06- | ···················· | ···················· | ····················· | ···················· | ···················· | ···················· |
| V | 5597 | hCoV-19/Brazil/SP-702/2020\|EPI_ISL_693242\|2020-06- | ···················· | ···················· | ····················· | ···················· | ···················· | ···················· |
| V | 5598 | hCoV-19/Brazil/SP-710/2020\|EPI_ISL_693243\|2020-06- | ···················· | ···················· | ····················· | ···················· | ···················· | ···················· |
| V | 5599 | hCoV-19/Australia/QLD1264/2020\|EPI_ISL_693266\|2020 | ···················· | ···················· | ····················· | ···················· | ···················· | ···················· |
| V | 5600 | hCoV-19/Australia/QLD1265/2020\|EPI_ISL_693267\|2020 | ···················· | ···················· | ····················· | ···················· | ···················· | ···················· |
| V | 5601 | hCoV-19/Australia/QLD1266/2020\|EPI_ISL_693268\|2020 | ···················· | ···················· | ····················· | ···················· | ···················· | ···················· |
| V | 5602 | hCoV-19/Australia/QLD1267/2020\|EPI_ISL_693269\|2020 | ···················· | ···················· | ····················· | ···················· | ···················· | ···················· |
| V | 5603 | hCoV-19/Australia/QLD1268/2020\|EPI_ISL_693270\|2020 | ···················· | ···················· | ····················· | ···················· | ···················· | ···················· |
| V | 5604 | hCoV-19/Australia/QLD1269/2020\|EPI_ISL_693271\|2020 | ···················· | ···················· | ····················· | ···················· | ···················· | ···················· |
| V | 5605 | hCoV-19/Australia/QLD1270/2020\|EPI_ISL_693272\|2020 | ···················· | ···················· | ····················· | ···················· | ···················· | ···················· |
| V | 5606 | hCoV-19/Australia/QLD1271/2020\|EPI_ISL_693273\|2020 | ···················· | ···················· | ····················· | ···················· | ···················· | ···················· |
| V | 5607 | hCoV-19/Australia/QLD1272/2020\|EPI_ISL_693274\|2020 | ···················· | ···················· | ····················· | ···················· | ···················· | ···················· |
| V | 5608 | hCoV-19/Australia/QLD1273/2020\|EPI_ISL_693275\|2020 | ···················· | ···················· | ····················· | ···················· | ···················· | ···················· |
| V | 5609 | hCoV-19/Australia/QLD1274/2020\|EPI_ISL_693276\|2020 | ···················· | ···················· | ····················· | ···················· | ···················· | ···················· |
| V | 5610 | hCoV-19/Australia/QLD1276/2020\|EPI_ISL_693277\|2020 | ···················· | ···················· | ····················· | ···················· | ···················· | ···················· |
| V | 5611 | hCoV-19/Australia/QLD1277/2020\|EPI_ISL_693278\|2020 | ···················· | ···················· | ····················· | ···················· | ···················· | ···················· |
| V | 5612 | hCoV-19/Australia/QLD1278/2020\|EPI_ISL_693279\|2020 | ···················· | ···················· | ····················· | ···················· | ······T············· | ···················· |
| V | 5613 | hCoV-19/Australia/QLD1279/2020\|EPI_ISL_693280\|2020 | ···················· | ···················· | ····················· | ···················· | ···················· | ···················· |
| V | 5614 | hCoV-19/Australia/QLD1280/2020\|EPI_ISL_693281\|2020 | ···················· | ···················· | ····················· | ···················· | ···················· | ···················· |
| V | 5615 | hCoV-19/Australia/QLD1282/2020\|EPI_ISL_693282\|2020 | ···················· | ···················· | ····················· | ···················· | ···················· | ···················· |
| V | 5616 | hCoV-19/Australia/QLD1283/2020\|EPI_ISL_693283\|2020 | ···················· | ···················· | ····················· | ···················· | ···················· | ···················· |
| V | 5617 | hCoV-19/Australia/QLD1285/2020\|EPI_ISL_693284\|2020 | ···················· | ···················· | ····················· | ···················· | ···················· | ···················· |
| V | 5618 | hCoV-19/Australia/QLD1286/2020\|EPI_ISL_693285\|2020 | ···················· | ···················· | ····················· | ···················· | ···················· | ···················· |
| V | 5619 | hCoV-19/Australia/QLD1287/2020\|EPI_ISL_693286\|2020 | ···················· | ···················· | ····················· | ···················· | ···················· | ···················· |
| V | 5620 | hCoV-19/Australia/QLD1288/2020\|EPI_ISL_693287\|2020 | ···················· | ···················· | ····················· | ···················· | ···················· | ···················· |
| V | 5621 | hCoV-19/Australia/QLD1289/2020\|EPI_ISL_693288\|2020 | ···················· | ···················· | ····················· | ···················· | ···················· | ···················· |
| V | 5622 | hCoV-19/Australia/QLD1292/2020\|EPI_ISL_693289\|2020 | ···················· | ···················· | ····················· | ···················· | ···················· | ···················· |
| V | 5623 | hCoV-19/Australia/QLD1293/2020\|EPI_ISL_693290\|2020 | ···················· | ···················· | ····················· | ···················· | ···················· | ···················· |
| V | 5624 | hCoV-19/Australia/QLD1296/2020\|EPI_ISL_693291\|2020 | ···················· | ···················· | ····················· | ···················· | ···················· | ···················· |
| V | 5625 | hCoV-19/Australia/QLD1297/2020\|EPI_ISL_693292\|2020 | ···················· | ···················· | ····················· | ···················· | ···················· | ···················· |
| V | 5626 | hCoV-19/Australia/QLD1299/2020\|EPI_ISL_693293\|2020 | ···················· | ···················· | ····················· | ···················· | ···················· | ···················· |
| V | 5627 | hCoV-19/Australia/QLD1300/2020\|EPI_ISL_693294\|2020 | ···················· | ···················· | ····················· | ···················· | ···················· | ···················· |
| V | 5628 | hCoV-19/Australia/QLD1302/2020\|EPI_ISL_693295\|2020 | ···················· | ···················· | ····················· | ···················· | ···················· | ···················· |
| V | 5629 | hCoV-19/Australia/QLD1303/2020\|EPI_ISL_693296\|2020 | ···················· | ···················· | ····················· | ···················· | ···················· | ···················· |
| V | 5630 | hCoV-19/Australia/QLD1304/2020\|EPI_ISL_693297\|2020 | ···················· | ···················· | ····················· | ···················· | ···················· | ···················· |
| V | 5631 | hCoV-19/Japan/YCU01/2020\|EPI_ISL_693298\|2020-07-29 | ···················· | ···················· | ····················· | ···················· | ···················· | ···················· |
| V | 5632 | hCoV-19/Japan/YCU02/2020\|EPI_ISL_693299\|2020-09-11 | ···················· | ···················· | ····················· | ···················· | ···················· | ···················· |
| V | 5633 | hCoV-19/Japan/PG-2395/2020\|EPI_ISL_686000\|2020-04 | ···················· | ···················· | ····················· | ···················· | ···················· | ···················· |
| V | 5634 | hCoV-19/Japan/PG-2396/2020\|EPI_ISL_686001\|2020-04 | ···················· | ···················· | ····················· | ···················· | ···················· | ···················· |
| V | 5635 | hCoV-19/Japan/PG-2397/2020\|EPI_ISL_686002\|2020-04 | ···················· | ···················· | ····················· | ···················· | ···················· | ···················· |
| V | 5636 | hCoV-19/Japan/PG-2398/2020\|EPI_ISL_686003\|2020-04 | ···················· | ···················· | ····················· | ···················· | ···················· | ···················· |
| V | 5637 | hCoV-19/Japan/PG-2399/2020\|EPI_ISL_686004\|2020-04 | ···················· | ···················· | ····················· | ···················· | ···················· | ···················· |
| V | 5638 | hCoV-19/Japan/PG-2400/2020\|EPI_ISL_686005\|2020-04 | ···················· | ···················· | ····················· | ···················· | ···················· | ···················· |
| V | 5639 | hCoV-19/Japan/PG-2401/2020\|EPI_ISL_686006\|2020-04 | ···················· | ···················· | ····················· | ···················· | ···················· | ···················· |
| V | 5640 | hCoV-19/Japan/PG-2402/2020\|EPI_ISL_686007\|2020-04 | ···················· | ···················· | ····················· | ···················· | ···················· | ···················· |
| V | 5641 | hCoV-19/Japan/PG-2403/2020\|EPI_ISL_686008\|2020-04 | ···················· | ···················· | ····················· | ···················· | ···················· | ···················· |
| V | 5642 | hCoV-19/Japan/PG-2404/2020\|EPI_ISL_686009\|2020-04 | ···················· | ···················· | ····················· | ···················· | ···················· | ···················· |
| V | 5643 | hCoV-19/Japan/PG-2405/2020\|EPI_ISL_686010\|2020-04 | ···················· | ···················· | ····················· | ···················· | ···················· | ···················· |
| V | 5644 | hCoV-19/Japan/PG-2406/2020\|EPI_ISL_686011\|2020-04 | ···················· | ···················· | ····················· | ···················· | ···················· | ···················· |
| V | 5645 | hCoV-19/Japan/PG-2407/2020\|EPI_ISL_686012\|2020-04 | ···················· | ···················· | ····················· | ···················· | ···················· | ···················· |
| V | 5646 | hCoV-19/Japan/PG-2408/2020\|EPI_ISL_686013\|2020-04 | ···················· | ···················· | ····················· | ···················· | ···················· | ···················· |
| V | 5647 | hCoV-19/Japan/PG-2409/2020\|EPI_ISL_686014\|2020-04 | ···················· | ···················· | ····················· | ···················· | ···················· | ···················· |

| | | | | | | | |
|---|---|---|---|---|---|---|---|
| V | 5648 hCoV-19/Japan/PG-2410/2020\|EPI_ISL_686015\|2020-04 | .................... | .................... | ..................... | .................... | .................... | .................... |
| V | 5649 hCoV-19/Japan/PG-2411/2020\|EPI_ISL_686016\|2020-04 | .................... | .................... | ..................... | .................... | .................... | .................... |
| V | 5650 hCoV-19/Japan/PG-2412/2020\|EPI_ISL_686017\|2020-04 | .................... | .................... | ..................... | .................... | .................... | .................... |
| V | 5651 hCoV-19/Japan/PG-2413/2020\|EPI_ISL_686018\|2020-04 | .................... | .................... | ..................... | .................... | .................... | .................... |
| V | 5652 hCoV-19/Japan/PG-2414/2020\|EPI_ISL_686019\|2020-04 | .................... | .................... | ..................... | .................... | .................... | .................... |
| V | 5653 hCoV-19/Japan/PG-2415/2020\|EPI_ISL_686020\|2020-04 | .................... | .................... | ..................... | .................... | .................... | .................... |
| V | 5654 hCoV-19/Japan/PG-2416/2020\|EPI_ISL_686021\|2020-03 | .................... | .................... | ..................... | .................... | .................... | .................... |
| V | 5655 hCoV-19/Japan/PG-2417/2020\|EPI_ISL_686022\|2020-03 | .................... | .................... | ..................... | .................... | .................... | .................... |
| V | 5656 hCoV-19/Japan/PG-2418/2020\|EPI_ISL_686023\|2020-03 | .................... | .................... | ..................... | .................... | .................... | .................... |
| V | 5657 hCoV-19/Japan/PG-2419/2020\|EPI_ISL_686024\|2020-03 | .................... | .................... | ..................... | .................... | .................... | .................... |
| V | 5658 hCoV-19/Japan/PG-2420/2020\|EPI_ISL_686025\|2020-04 | .................... | .................... | ..................... | .................... | .................... | .................... |
| V | 5659 hCoV-19/Japan/PG-2421/2020\|EPI_ISL_686026\|2020-04 | .................... | .................... | ..................... | .................... | .................... | .................... |
| V | 5660 hCoV-19/Japan/PG-2422/2020\|EPI_ISL_686027\|2020-04 | .................... | .................... | ..................... | .................... | .................... | .................... |
| V | 5661 hCoV-19/Japan/PG-2423/2020\|EPI_ISL_686028\|2020-04 | .................... | .................... | ..................... | .................... | .................... | .................... |
| V | 5662 hCoV-19/Japan/PG-2424/2020\|EPI_ISL_686029\|2020-04 | .................... | .................... | ..................... | .................... | .................... | .................... |
| V | 5663 hCoV-19/Japan/PG-2425/2020\|EPI_ISL_686030\|2020-04 | .................... | .................... | ..................... | .................... | .................... | .................... |
| V | 5664 hCoV-19/Japan/PG-2426/2020\|EPI_ISL_686031\|2020-04 | .................... | .................... | ..................... | .................... | .................... | .................... |
| V | 5665 hCoV-19/Japan/PG-2427/2020\|EPI_ISL_686032\|2020-04 | .................... | .................... | ..................... | .................... | .................... | .................... |
| V | 5666 hCoV-19/Japan/PG-2428/2020\|EPI_ISL_686033\|2020-04 | .................... | .................... | ..................... | .................... | .................... | .................... |
| V | 5667 hCoV-19/Japan/PG-2429/2020\|EPI_ISL_686034\|2020-04 | .................... | .................... | ..................... | .................... | .................... | .................... |
| V | 5668 hCoV-19/Japan/PG-2430/2020\|EPI_ISL_686035\|2020-04 | .................... | .................... | ..................... | .................... | .................... | .................... |
| V | 5669 hCoV-19/Japan/PG-2431/2020\|EPI_ISL_686036\|2020-04 | .................... | .................... | ..................... | .................... | .................... | .................... |
| V | 5670 hCoV-19/Japan/PG-2432/2020\|EPI_ISL_686037\|2020-04 | .................... | .................... | ..................... | .................... | .................... | .................... |
| V | 5671 hCoV-19/Japan/PG-2433/2020\|EPI_ISL_686038\|2020-03 | .................... | .................... | ..................... | .................... | .................... | .................... |
| V | 5672 hCoV-19/Japan/PG-2434/2020\|EPI_ISL_686039\|2020-03 | .................... | .................... | ..................... | .................... | .................... | .................... |
| V | 5673 hCoV-19/Japan/PG-2435/2020\|EPI_ISL_686040\|2020-03 | .................... | .................... | ..................... | .................... | .................... | .................... |
| V | 5674 hCoV-19/Japan/PG-2436/2020\|EPI_ISL_686041\|2020-03 | .................... | .................... | ..................... | .................... | .................... | .................... |
| V | 5675 hCoV-19/Japan/PG-2437/2020\|EPI_ISL_686042\|2020-03 | .................... | .................... | ..................... | .................... | .................... | .................... |
| V | 5676 hCoV-19/Japan/PG-2438/2020\|EPI_ISL_686043\|2020-03 | .................... | .................... | ..................... | .................... | .................... | .................... |
| V | 5677 hCoV-19/Japan/PG-2439/2020\|EPI_ISL_686044\|2020-04 | .................... | .................... | ..................... | .................... | .................... | .................... |
| V | 5678 hCoV-19/Japan/PG-2440/2020\|EPI_ISL_686045\|2020-04 | .................... | .................... | ..................... | .................... | .................... | .................... |
| V | 5679 hCoV-19/Japan/PG-2441/2020\|EPI_ISL_686046\|2020-04 | .................... | .................... | ..................... | .................... | .................... | .................... |
| V | 5680 hCoV-19/Japan/PG-2442/2020\|EPI_ISL_686047\|2020-04 | .................... | .................... | ..................... | .................... | .................... | .................... |
| V | 5681 hCoV-19/Japan/PG-2443/2020\|EPI_ISL_686048\|2020-04 | .................... | .................... | ..................... | .................... | .................... | .................... |
| V | 5682 hCoV-19/Japan/PG-2444/2020\|EPI_ISL_686049\|2020-04 | .................... | .................... | ..................... | .................... | .................... | .................... |
| V | 5683 hCoV-19/Japan/PG-2445/2020\|EPI_ISL_686050\|2020-04 | .................... | .................... | ..................... | .................... | .................... | .................... |
| V | 5684 hCoV-19/Japan/PG-2446/2020\|EPI_ISL_686051\|2020-04 | .................... | .................... | ..................... | .................... | .................... | .................... |
| V | 5685 hCoV-19/Japan/PG-2447/2020\|EPI_ISL_686052\|2020-04 | .................... | .................... | ..................... | .................... | .................... | .................... |
| V | 5686 hCoV-19/Japan/PG-2448/2020\|EPI_ISL_686053\|2020-04 | .................... | .................... | ..................... | .................... | .................... | .................... |
| V | 5687 hCoV-19/Japan/PG-2449/2020\|EPI_ISL_686054\|2020-04 | .................... | .................... | ..................... | .................... | .................... | .................... |
| V | 5688 hCoV-19/Japan/PG-2450/2020\|EPI_ISL_686055\|2020-04 | .................... | .................... | ..................... | .................... | .................... | .................... |
| V | 5689 hCoV-19/Japan/PG-2451/2020\|EPI_ISL_686056\|2020-04 | .................... | .................... | ..................... | .................... | .................... | .................... |
| V | 5690 hCoV-19/Japan/PG-2452/2020\|EPI_ISL_686057\|2020-04 | .................... | .................... | ..................... | .................... | .................... | .................... |
| V | 5691 hCoV-19/Japan/PG-2453/2020\|EPI_ISL_686058\|2020-04 | .................... | .................... | ..................... | .................... | .................... | .................... |
| V | 5692 hCoV-19/Japan/PG-2454/2020\|EPI_ISL_686059\|2020-04 | .................... | .................... | ..................... | .................... | .................... | .................... |
| V | 5693 hCoV-19/Japan/PG-8205/2020\|EPI_ISL_687304\|2020-08 | .................... | .................... | ..................... | .................... | .................... | .................... |
| V | 5694 hCoV-19/Japan/PG-8206/2020\|EPI_ISL_687305\|2020-08 | .................... | .................... | ..................... | .................... | .................... | .................... |
| V | 5695 hCoV-19/Japan/PG-8207/2020\|EPI_ISL_687306\|2020-08 | .................... | .................... | ..................... | .................... | .................... | .................... |
| V | 5696 hCoV-19/Japan/PG-8208/2020\|EPI_ISL_687307\|2020-08 | .................... | .................... | ..................... | .................... | .................... | .................... |
| V | 5697 hCoV-19/Japan/PG-8209/2020\|EPI_ISL_687308\|2020-08 | .................... | .................... | ..................... | .................... | .................... | .................... |
| V | 5698 hCoV-19/Japan/PG-8210/2020\|EPI_ISL_687309\|2020-08 | .................... | .................... | ..................... | .................... | .................... | .................... |
| V | 5699 hCoV-19/Japan/PG-8211/2020\|EPI_ISL_687310\|2020-08 | .................... | .................... | ..................... | .................... | .................... | .................... |
| V | 5700 hCoV-19/Japan/PG-8212/2020\|EPI_ISL_687311\|2020-08 | .................... | .................... | ..................... | .................... | .................... | .................... |
| V | 5701 hCoV-19/Japan/PG-8213/2020\|EPI_ISL_687312\|2020-08 | .................... | .................... | ..................... | .................... | .................... | .................... |
| V | 5702 hCoV-19/Japan/PG-8214/2020\|EPI_ISL_687313\|2020-08 | .................... | .................... | ..................... | .................... | .................... | .................... |
| V | 5703 hCoV-19/Japan/PG-8215/2020\|EPI_ISL_687314\|2020-08 | .................... | .................... | ..................... | .................... | .................... | .................... |
| V | 5704 hCoV-19/Japan/PG-8216/2020\|EPI_ISL_687315\|2020-08 | .................... | .................... | ..................... | .................... | .................... | .................... |
| V | 5705 hCoV-19/Japan/PG-8217/2020\|EPI_ISL_687316\|2020-08 | .................... | .................... | ..................... | .................... | .................... | .................... |
| V | 5706 hCoV-19/Japan/PG-8218/2020\|EPI_ISL_687317\|2020-08 | .................... | .................... | ..................... | .................... | .................... | .................... |
| V | 5707 hCoV-19/Japan/PG-8219/2020\|EPI_ISL_687318\|2020-08 | .................... | .................... | ..................... | .................... | .................... | .................... |
| V | 5708 hCoV-19/Japan/PG-8220/2020\|EPI_ISL_687319\|2020-08 | .................... | .................... | ..................... | .................... | .................... | .................... |
| V | 5709 hCoV-19/Japan/PG-8221/2020\|EPI_ISL_687320\|2020-08 | .................... | .................... | ..................... | .................... | .................... | .................... |
| V | 5710 hCoV-19/Japan/PG-8222/2020\|EPI_ISL_687321\|2020-08 | .................... | .................... | ..................... | .................... | .................... | .................... |
| V | 5711 hCoV-19/Japan/PG-8223/2020\|EPI_ISL_687322\|2020-08 | .................... | .................... | ..................... | .................... | .................... | .................... |
| V | 5712 hCoV-19/Japan/PG-8224/2020\|EPI_ISL_687323\|2020-08 | .................... | .................... | ..................... | .................... | .................... | .................... |
| V | 5713 hCoV-19/Japan/PG-8225/2020\|EPI_ISL_687324\|2020-08 | .................... | .................... | ..................... | .................... | .................... | .................... |
| V | 5714 hCoV-19/Japan/PG-8226/2020\|EPI_ISL_687325\|2020-08 | .................... | .................... | ..................... | .................... | .................... | .................... |
| V | 5715 hCoV-19/Japan/PG-8227/2020\|EPI_ISL_687326\|2020-08 | .................... | .................... | ..................... | .................... | .................... | .................... |
| V | 5716 hCoV-19/Japan/PG-8228/2020\|EPI_ISL_687327\|2020-08 | .................... | .................... | ..................... | .................... | .................... | .................... |
| V | 5717 hCoV-19/Japan/PG-8229/2020\|EPI_ISL_687328\|2020-08 | .................... | .................... | ..................... | .................... | .................... | .................... |
| V | 5718 hCoV-19/Japan/PG-8230/2020\|EPI_ISL_687329\|2020-08 | .................... | .................... | ..................... | .................... | .................... | .................... |
| V | 5719 hCoV-19/Japan/PG-8231/2020\|EPI_ISL_687330\|2020-08 | .................... | .................... | ..................... | .................... | .................... | .................... |
| V | 5720 hCoV-19/Japan/PG-8232/2020\|EPI_ISL_687331\|2020-08 | .................... | .................... | ..................... | .................... | .................... | .................... |
| V | 5721 hCoV-19/Japan/PG-8233/2020\|EPI_ISL_687332\|2020-08 | .................... | .................... | ..................... | .................... | .................... | .................... |
| V | 5722 hCoV-19/Japan/PG-8234/2020\|EPI_ISL_687333\|2020-08 | .................... | .................... | ..................... | .................... | .................... | .................... |
| V | 5723 hCoV-19/Japan/PG-8235/2020\|EPI_ISL_687334\|2020-08 | .................... | .................... | ..................... | .................... | .................... | .................... |
| V | 5724 hCoV-19/Japan/PG-8236/2020\|EPI_ISL_687335\|2020-08 | .................... | .................... | ..................... | .................... | .................... | .................... |
| V | 5725 hCoV-19/Japan/PG-8237/2020\|EPI_ISL_687336\|2020-08 | .................... | .................... | ..................... | .................... | .................... | .................... |
| V | 5726 hCoV-19/Japan/PG-8238/2020\|EPI_ISL_687337\|2020-08 | .................... | .................... | ..................... | .................... | .................... | .................... |
| V | 5727 hCoV-19/Japan/PG-8239/2020\|EPI_ISL_687338\|2020-08 | .................... | .................... | ..................... | .................... | .................... | .................... |

| | | | | | | | | |
|---|---|---|---|---|---|---|---|---|
| V | 5728 | hCoV-19/Japan/PG-8240/2020\|EPI_ISL_687339\|2020-08 | .................... | .................... | ..................... | .................... | .................... | .................... |
| V | 5729 | hCoV-19/Japan/PG-8241/2020\|EPI_ISL_687340\|2020-08 | .................... | .................... | ..................... | .................... | .................... | .................... |
| V | 5730 | hCoV-19/Japan/PG-8242/2020\|EPI_ISL_687341\|2020-08 | .................... | .................... | ..................... | .................... | .................... | .................... |
| V | 5731 | hCoV-19/Japan/PG-8243/2020\|EPI_ISL_687342\|2020-08 | .................... | .................... | ..................... | .................... | .................... | .................... |
| V | 5733 | hCoV-19/Japan/PG-8245/2020\|EPI_ISL_687344\|2020-08 | .................... | .................... | ..................... | .................... | .................... | .................... |
| V | 5734 | hCoV-19/Japan/PG-8246/2020\|EPI_ISL_687345\|2020-08 | .................... | .................... | ..................... | .................... | .................... | .................... |
| V | 5735 | hCoV-19/Japan/PG-8247/2020\|EPI_ISL_687346\|2020-08 | .................... | .................... | ..................... | .................... | .................... | .................... |
| V | 5736 | hCoV-19/Japan/PG-8248/2020\|EPI_ISL_687347\|2020-08 | .................... | .................... | ..................... | .................... | .................... | .................... |
| V | 5737 | hCoV-19/Japan/PG-8249/2020\|EPI_ISL_687348\|2020-08 | .................... | .................... | ..................... | .................... | .................... | .................... |
| V | 5738 | hCoV-19/Japan/PG-8250/2020\|EPI_ISL_687349\|2020-08 | .................... | .................... | ..................... | .................... | .................... | .................... |
| V | 5739 | hCoV-19/Japan/PG-8251/2020\|EPI_ISL_687350\|2020-08 | .................... | .................... | ..................... | .................... | .................... | .................... |
| V | 5740 | hCoV-19/Japan/PG-8252/2020\|EPI_ISL_687351\|2020-08 | .................... | .................... | ..................... | .................... | .................... | .................... |
| V | 5741 | hCoV-19/Japan/PG-8253/2020\|EPI_ISL_687352\|2020-08 | .................... | .................... | ..................... | .................... | .................... | .................... |
| V | 5742 | hCoV-19/Japan/PG-8254/2020\|EPI_ISL_687353\|2020-08 | .................... | .................... | ..................... | .................... | .................... | .................... |
| V | 5743 | hCoV-19/Japan/PG-8255/2020\|EPI_ISL_687354\|2020-08 | .................... | .................... | ..................... | .................... | .................... | .................... |
| V | 5744 | hCoV-19/Japan/PG-8256/2020\|EPI_ISL_687355\|2020-08 | .................... | .................... | ..................... | .................... | .................... | .................... |
| V | 5745 | hCoV-19/Japan/PG-8257/2020\|EPI_ISL_687356\|2020-08 | .................... | .................... | ..................... | .................... | .................... | .................... |
| V | 5746 | hCoV-19/Japan/PG-8258/2020\|EPI_ISL_687357\|2020-08 | .................... | .................... | ..................... | .................... | .................... | .................... |
| V | 5747 | hCoV-19/Japan/PG-8259/2020\|EPI_ISL_687358\|2020-08 | .................... | .................... | ..................... | .................... | .................... | .................... |
| V | 5748 | hCoV-19/Japan/PG-8260/2020\|EPI_ISL_687359\|2020-08 | .................... | .................... | ..................... | .................... | .................... | .................... |
| V | 5749 | hCoV-19/Japan/PG-8261/2020\|EPI_ISL_687360\|2020-09 | .................... | .................... | ..................... | .................... | .................... | .................... |
| V | 5750 | hCoV-19/Japan/PG-8262/2020\|EPI_ISL_687361\|2020-08 | .................... | .................... | ..................... | .................... | .................... | .................... |
| V | 5751 | hCoV-19/Japan/PG-8263/2020\|EPI_ISL_687362\|2020-08 | .................... | .................... | ..................... | .................... | .................... | .................... |
| V | 5752 | hCoV-19/Japan/PG-8264/2020\|EPI_ISL_687363\|2020-08 | .................... | .................... | ..................... | .................... | .................... | .................... |
| V | 5753 | hCoV-19/Japan/PG-8265/2020\|EPI_ISL_687364\|2020-08 | .................... | .................... | ..................... | .................... | .................... | .................... |
| V | 5754 | hCoV-19/Japan/PG-8266/2020\|EPI_ISL_687365\|2020-08 | .................... | .................... | ..................... | .................... | .................... | .................... |
| V | 5755 | hCoV-19/Japan/PG-8267/2020\|EPI_ISL_687366\|2020-08 | .................... | .................... | ..................... | .................... | .................... | .................... |
| V | 5756 | hCoV-19/Japan/PG-8268/2020\|EPI_ISL_687367\|2020-08 | .................... | .................... | ..................... | .................... | .................... | .................... |
| V | 5757 | hCoV-19/Japan/PG-8269/2020\|EPI_ISL_687368\|2020-08 | .................... | .................... | ..................... | .................... | .................... | .................... |
| V | 5758 | hCoV-19/Japan/PG-8270/2020\|EPI_ISL_687369\|2020-08 | .................... | .................... | ..................... | .................... | .................... | .................... |
| V | 5759 | hCoV-19/Japan/PG-8271/2020\|EPI_ISL_687370\|2020-08 | .................... | .................... | ..................... | .................... | .................... | .................... |
| V | 5760 | hCoV-19/Japan/PG-8272/2020\|EPI_ISL_687371\|2020-08 | .................... | .................... | ..................... | .................... | .................... | .................... |
| V | 5761 | hCoV-19/Japan/PG-8273/2020\|EPI_ISL_687372\|2020-08 | .................... | .................... | ..................... | .................... | .................... | .................... |
| V | 5762 | hCoV-19/Japan/PG-8274/2020\|EPI_ISL_687373\|2020-08 | .................... | .................... | ..................... | .................... | .................... | .................... |
| V | 5763 | hCoV-19/Japan/PG-8275/2020\|EPI_ISL_687374\|2020-08 | .................... | .................... | ..................... | .................... | .................... | .................... |
| V | 5764 | hCoV-19/Japan/PG-8276/2020\|EPI_ISL_687375\|2020-08 | .................... | .................... | ..................... | .................... | .................... | .................... |
| V | 5765 | hCoV-19/Japan/PG-8277/2020\|EPI_ISL_687376\|2020-08 | .................... | .................... | ..................... | .................... | .................... | .................... |
| V | 5766 | hCoV-19/Japan/PG-8278/2020\|EPI_ISL_687377\|2020-08 | .................... | .................... | ..................... | .................... | .................... | .................... |
| V | 5767 | hCoV-19/Japan/PG-8279/2020\|EPI_ISL_687378\|2020-08 | .................... | .................... | ..................... | .................... | .................... | .................... |
| V | 5768 | hCoV-19/Japan/PG-8280/2020\|EPI_ISL_687379\|2020-08 | .................... | .................... | ..................... | .................... | .................... | .................... |
| V | 5769 | hCoV-19/Japan/PG-8281/2020\|EPI_ISL_687380\|2020-08 | .................... | .................... | ..................... | .................... | .................... | .................... |
| V | 5770 | hCoV-19/Japan/PG-8282/2020\|EPI_ISL_687381\|2020-08 | .................... | .................... | ..................... | .................... | .................... | .................... |
| V | 5771 | hCoV-19/Japan/PG-8283/2020\|EPI_ISL_687382\|2020-08 | .................... | .................... | ..................... | .................... | .................... | .................... |
| V | 5772 | hCoV-19/Japan/PG-8284/2020\|EPI_ISL_687383\|2020-08 | .................... | .................... | ..................... | .................... | .................... | .................... |
| V | 5773 | hCoV-19/Japan/PG-8285/2020\|EPI_ISL_687384\|2020-08 | .................... | .................... | ..................... | .................... | .................... | .................... |
| V | 5774 | hCoV-19/Japan/PG-8286/2020\|EPI_ISL_687385\|2020-08 | .................... | .................... | ..................... | .................... | .................... | .................... |
| V | 5775 | hCoV-19/Japan/PG-8287/2020\|EPI_ISL_687386\|2020-08 | .................... | .................... | ..................... | .................... | .................... | .................... |
| V | 5776 | hCoV-19/Japan/PG-8288/2020\|EPI_ISL_687387\|2020-08 | .................... | .................... | ..................... | .................... | .................... | .................... |
| V | 5777 | hCoV-19/Japan/PG-8289/2020\|EPI_ISL_687388\|2020-08 | .................... | .................... | ..................... | .................... | .................... | .................... |
| V | 5778 | hCoV-19/Japan/PG-8290/2020\|EPI_ISL_687389\|2020-08 | .................... | .................... | ..................... | .................... | .................... | .................... |
| V | 5779 | hCoV-19/Japan/PG-5510/2020\|EPI_ISL_688635\|2020-08 | .................... | .................... | ..................... | .................... | .................... | .................... |
| V | 5780 | hCoV-19/Japan/PG-5511/2020\|EPI_ISL_688636\|2020-08 | .................... | .................... | ..................... | .................... | .................... | .................... |
| V | 5781 | hCoV-19/Japan/PG-5512/2020\|EPI_ISL_688637\|2020-08 | .................... | .................... | ..................... | .................... | .................... | .................... |
| V | 5782 | hCoV-19/Japan/PG-5513/2020\|EPI_ISL_688638\|2020-08 | .................... | .................... | ..................... | .................... | .................... | .................... |
| V | 5783 | hCoV-19/Japan/PG-5514/2020\|EPI_ISL_688639\|2020-08 | .................... | .................... | ..................... | .................... | .................... | .................... |
| V | 5784 | hCoV-19/Japan/PG-5515/2020\|EPI_ISL_688640\|2020-08 | .................... | .................... | ..................... | .................... | .................... | .................... |
| V | 5785 | hCoV-19/Japan/PG-5516/2020\|EPI_ISL_688641\|2020-08 | .................... | .................... | ..................... | .................... | .................... | .................... |
| V | 5786 | hCoV-19/Japan/PG-5517/2020\|EPI_ISL_688642\|2020-08 | .................... | .................... | ..................... | .................... | .................... | .................... |
| V | 5787 | hCoV-19/Japan/PG-5518/2020\|EPI_ISL_688643\|2020-08 | .................... | .................... | ..................... | .................... | .................... | .................... |
| V | 5788 | hCoV-19/Japan/PG-5519/2020\|EPI_ISL_688644\|2020-08 | .................... | .................... | ..................... | .................... | .................... | .................... |
| V | 5789 | hCoV-19/Japan/PG-5520/2020\|EPI_ISL_688645\|2020-08 | .................... | .................... | ..................... | .................... | .................... | .................... |
| V | 5790 | hCoV-19/Japan/PG-5521/2020\|EPI_ISL_688646\|2020-08 | .................... | .................... | ..................... | .................... | .................... | .................... |
| V | 5791 | hCoV-19/Japan/PG-5522/2020\|EPI_ISL_688647\|2020-08 | .................... | .................... | ..................... | .................... | .................... | .................... |
| V | 5792 | hCoV-19/Japan/PG-5523/2020\|EPI_ISL_688648\|2020-08 | .................... | .................... | ..................... | .................... | .................... | .................... |
| V | 5793 | hCoV-19/Japan/PG-5524/2020\|EPI_ISL_688649\|2020-08 | .................... | .................... | ..................... | .................... | .................... | .................... |
| V | 5794 | hCoV-19/Japan/PG-5525/2020\|EPI_ISL_688650\|2020-08 | .................... | .................... | ..................... | .................... | .................... | .................... |
| V | 5795 | hCoV-19/Japan/PG-5526/2020\|EPI_ISL_688651\|2020-08 | .................... | .................... | ..................... | .................... | .................... | .................... |
| V | 5796 | hCoV-19/Japan/PG-5527/2020\|EPI_ISL_688652\|2020-08 | .................... | .................... | ..................... | .................... | .................... | .................... |
| V | 5797 | hCoV-19/Japan/PG-5528/2020\|EPI_ISL_688653\|2020-08 | .................... | .................... | ..................... | .................... | .................... | .................... |
| V | 5798 | hCoV-19/Japan/PG-5529/2020\|EPI_ISL_688654\|2020-08 | .................... | .................... | ..................... | .................... | .................... | .................... |
| V | 5799 | hCoV-19/Japan/PG-5530/2020\|EPI_ISL_688655\|2020-08 | .................... | .................... | ..................... | .................... | .................... | .................... |
| V | 5800 | hCoV-19/Japan/PG-5531/2020\|EPI_ISL_688656\|2020-08 | .................... | .................... | ..................... | .................... | .................... | .................... |
| V | 5801 | hCoV-19/Japan/PG-5532/2020\|EPI_ISL_688657\|2020-08 | .................... | .................... | ..................... | .................... | .................... | .................... |
| V | 5802 | hCoV-19/Japan/PG-5533/2020\|EPI_ISL_688658\|2020-08 | .................... | .................... | ..................... | .................... | .................... | .................... |
| V | 5803 | hCoV-19/Japan/PG-5534/2020\|EPI_ISL_688659\|2020-08 | .................... | .................... | ..................... | .................... | .................... | .................... |
| V | 5804 | hCoV-19/Japan/PG-5535/2020\|EPI_ISL_688660\|2020-08 | .................... | .................... | ..................... | .................... | .................... | .................... |
| V | 5805 | hCoV-19/Japan/PG-5536/2020\|EPI_ISL_688661\|2020-08 | .................... | .................... | ..................... | .................... | .................... | .................... |
| V | 5806 | hCoV-19/Japan/PG-5537/2020\|EPI_ISL_688662\|2020-08 | .................... | .................... | ..................... | .................... | .................... | .................... |
| V | 5807 | hCoV-19/Japan/PG-5538/2020\|EPI_ISL_688663\|2020-08 | .................... | .................... | ..................... | .................... | .................... | .................... |
| V | 5808 | hCoV-19/Japan/PG-5539/2020\|EPI_ISL_688664\|2020-08 | .................... | .................... | ..................... | .................... | .................... | .................... |

| | | | | | | | |
|---|---|---|---|---|---|---|---|
| V | 5809 hCoV-19/Japan/PG-5540/2020\|EPI_ISL_688665\|2020-08 | ···················· | ···················· | ····················· | ···················· | ···················· | ···················· |
| V | 5810 hCoV-19/Japan/PG-5541/2020\|EPI_ISL_688666\|2020-08 | ···················· | ···················· | ····················· | ···················· | ···················· | ···················· |
| V | 5811 hCoV-19/Japan/PG-5542/2020\|EPI_ISL_688667\|2020-08 | ···················· | ···················· | ····················· | ···················· | ···················· | ···················· |
| V | 5812 hCoV-19/Japan/PG-5543/2020\|EPI_ISL_688668\|2020-08 | ···················· | ···················· | ····················· | ···················· | ···················· | ···················· |
| V | 5813 hCoV-19/Japan/PG-5544/2020\|EPI_ISL_688669\|2020-08 | ···················· | ···················· | ····················· | ···················· | ···················· | ···················· |
| V | 5814 hCoV-19/Japan/PG-5545/2020\|EPI_ISL_688670\|2020-08 | ···················· | ···················· | ····················· | ···················· | ···················· | ···················· |
| V | 5815 hCoV-19/Japan/PG-5546/2020\|EPI_ISL_688671\|2020-08 | ···················· | ···················· | ····················· | ···················· | ···················· | ···················· |
| V | 5816 hCoV-19/Japan/PG-5547/2020\|EPI_ISL_688672\|2020-08 | ···················· | ···················· | ····················· | ···················· | ···················· | ···················· |
| V | 5817 hCoV-19/Japan/PG-5548/2020\|EPI_ISL_688673\|2020-08 | ···················· | ···················· | ····················· | ···················· | ···················· | ···················· |
| V | 5818 hCoV-19/Japan/PG-5549/2020\|EPI_ISL_688674\|2020-08 | ···················· | ···················· | ····················· | ···················· | ···················· | ···················· |
| V | 5819 hCoV-19/Japan/PG-5550/2020\|EPI_ISL_688675\|2020-08 | ···················· | ···················· | ····················· | ···················· | ···················· | ···················· |
| V | 5820 hCoV-19/Japan/PG-5551/2020\|EPI_ISL_688676\|2020-08 | ···················· | ···················· | ····················· | ···················· | ···················· | ···················· |
| V | 5821 hCoV-19/Japan/PG-5552/2020\|EPI_ISL_688677\|2020-08 | ···················· | ···················· | ····················· | ···················· | ···················· | ···················· |
| V | 5822 hCoV-19/Japan/PG-5553/2020\|EPI_ISL_688678\|2020-08 | ···················· | ···················· | ····················· | ···················· | ···················· | ···················· |
| V | 5823 hCoV-19/Japan/PG-5554/2020\|EPI_ISL_688679\|2020-08 | ···················· | ···················· | ····················· | ···················· | ···················· | ···················· |
| V | 5824 hCoV-19/Japan/PG-5555/2020\|EPI_ISL_688680\|2020-08 | ···················· | ···················· | ····················· | ···················· | ···················· | ···················· |
| V | 5825 hCoV-19/Japan/PG-5556/2020\|EPI_ISL_688681\|2020-08 | ···················· | ···················· | ····················· | ···················· | ···················· | ···················· |
| V | 5826 hCoV-19/Japan/PG-5557/2020\|EPI_ISL_688682\|2020-08 | ···················· | ···················· | ····················· | ···················· | ···················· | ···················· |
| V | 5827 hCoV-19/Japan/PG-5558/2020\|EPI_ISL_688683\|2020-08 | ···················· | ···················· | ····················· | ···················· | ···················· | ···················· |
| V | 5828 hCoV-19/Japan/PG-5559/2020\|EPI_ISL_688684\|2020-08 | ···················· | ···················· | ····················· | ···················· | ···················· | ···················· |
| V | 5829 hCoV-19/Japan/PG-5560/2020\|EPI_ISL_688685\|2020-08 | ···················· | ···················· | ····················· | ···················· | ···················· | ···················· |
| V | 5830 hCoV-19/Japan/PG-5561/2020\|EPI_ISL_688686\|2020-08 | ···················· | ···················· | ····················· | ···················· | ···················· | ···················· |
| V | 5831 hCoV-19/Japan/PG-5562/2020\|EPI_ISL_688687\|2020-08 | ···················· | ···················· | ····················· | ···················· | ···················· | ···················· |
| V | 5832 hCoV-19/Japan/PG-5563/2020\|EPI_ISL_688688\|2020-08 | ···················· | ···················· | ····················· | ···················· | ···················· | ···················· |
| V | 5833 hCoV-19/Japan/PG-5564/2020\|EPI_ISL_688689\|2020-08 | ···················· | ···················· | ····················· | ···················· | ···················· | ···················· |
| V | 5834 hCoV-19/Japan/PG-5565/2020\|EPI_ISL_688690\|2020-08 | ···················· | ···················· | ····················· | ···················· | ···················· | ···················· |
| V | 5835 hCoV-19/Japan/PG-5566/2020\|EPI_ISL_688691\|2020-08 | ···················· | ···················· | ····················· | ···················· | ···················· | ···················· |
| V | 5836 hCoV-19/Japan/PG-5567/2020\|EPI_ISL_688692\|2020-08 | ···················· | ···················· | ····················· | ···················· | ···················· | ···················· |
| V | 5837 hCoV-19/Japan/PG-5568/2020\|EPI_ISL_688693\|2020-08 | ···················· | ···················· | ····················· | ···················· | ···················· | ···················· |
| V | 5839 hCoV-19/Japan/PG-5570/2020\|EPI_ISL_688695\|2020-08 | ···················· | ···················· | ····················· | ···················· | ···················· | ···················· |
| V | 5840 hCoV-19/Japan/PG-5571/2020\|EPI_ISL_688696\|2020-08 | ···················· | ···················· | ····················· | ···················· | ···················· | ···················· |
| V | 5841 hCoV-19/Japan/PG-5572/2020\|EPI_ISL_688697\|2020-08 | ···················· | ···················· | ····················· | ···················· | ···················· | ···················· |
| V | 5842 hCoV-19/Japan/PG-5573/2020\|EPI_ISL_688698\|2020-08 | ···················· | ···················· | ····················· | ···················· | ···················· | ···················· |
| V | 5843 hCoV-19/Japan/PG-5574/2020\|EPI_ISL_688699\|2020-08 | ···················· | ···················· | ····················· | ···················· | ···················· | ···················· |
| V | 5844 hCoV-19/Japan/PG-4844/2020\|EPI_ISL_689966\|2020-07 | ···················· | ···················· | ····················· | ···················· | ···················· | ···················· |
| V | 5845 hCoV-19/Japan/PG-4845/2020\|EPI_ISL_689967\|2020-07 | ···················· | ···················· | ····················· | ···················· | ···················· | ···················· |
| V | 5846 hCoV-19/Japan/PG-4846/2020\|EPI_ISL_689968\|2020-08 | ···················· | ···················· | ····················· | ···················· | ···················· | ···················· |
| V | 5847 hCoV-19/Japan/PG-4847/2020\|EPI_ISL_689969\|2020-08 | ···················· | ···················· | ····················· | ···················· | ···················· | ···················· |
| V | 5848 hCoV-19/Japan/PG-4848/2020\|EPI_ISL_689970\|2020-08 | ···················· | ···················· | ····················· | ···················· | ···················· | ···················· |
| V | 5849 hCoV-19/Japan/PG-4849/2020\|EPI_ISL_689971\|2020-07 | ···················· | ···················· | ····················· | ···················· | ···················· | ···················· |
| V | 5850 hCoV-19/Japan/PG-4850/2020\|EPI_ISL_689972\|2020-07 | ···················· | ···················· | ····················· | ···················· | ···················· | ···················· |
| V | 5851 hCoV-19/Japan/PG-4851/2020\|EPI_ISL_689973\|2020-07 | ···················· | ···················· | ····················· | ···················· | ···················· | ···················· |
| V | 5852 hCoV-19/Japan/PG-4852/2020\|EPI_ISL_689974\|2020-07 | ···················· | ···················· | ····················· | ···················· | ···················· | ···················· |
| V | 5853 hCoV-19/Japan/PG-4853/2020\|EPI_ISL_689975\|2020-07 | ···················· | ···················· | ····················· | ···················· | ···················· | ···················· |
| V | 5854 hCoV-19/Japan/PG-4854/2020\|EPI_ISL_689976\|2020-07 | ···················· | ···················· | ····················· | ···················· | ···················· | ···················· |
| V | 5855 hCoV-19/Japan/PG-4855/2020\|EPI_ISL_689977\|2020-07 | ···················· | ···················· | ····················· | ···················· | ···················· | ···················· |
| V | 5856 hCoV-19/Japan/PG-2823/2020\|EPI_ISL_690950\|2020-04 | ···················· | ···················· | ····················· | ···················· | ···················· | ···················· |
| V | 5857 hCoV-19/Japan/PG-2824/2020\|EPI_ISL_690951\|2020-04 | ···················· | ···················· | ····················· | ···················· | ···················· | ···················· |
| V | 5858 hCoV-19/Japan/PG-2825/2020\|EPI_ISL_690952\|2020-04 | ···················· | ···················· | ····················· | ···················· | ···················· | ···················· |
| V | 5859 hCoV-19/Japan/PG-2826/2020\|EPI_ISL_690953\|2020-04 | ···················· | ···················· | ····················· | ···················· | ···················· | ···················· |
| V | 5860 hCoV-19/Japan/PG-2827/2020\|EPI_ISL_690954\|2020-04 | ···················· | ···················· | ····················· | ···················· | ···················· | ···················· |
| V | 5861 hCoV-19/Japan/PG-2828/2020\|EPI_ISL_690955\|2020-04 | ···················· | ···················· | ····················· | ···················· | ···················· | ···················· |
| V | 5862 hCoV-19/Japan/PG-2829/2020\|EPI_ISL_690956\|2020-04 | ···················· | ···················· | ····················· | ···················· | ···················· | ···················· |
| V | 5863 hCoV-19/Japan/PG-2830/2020\|EPI_ISL_690957\|2020-04 | ···················· | ···················· | ····················· | ···················· | ···················· | ···················· |
| V | 5864 hCoV-19/Japan/PG-2831/2020\|EPI_ISL_690958\|2020-04 | ···················· | ···················· | ····················· | ···················· | ···················· | ···················· |
| V | 5865 hCoV-19/Japan/PG-2832/2020\|EPI_ISL_690959\|2020-04 | ···················· | ·A·················· | ····················· | ···················· | ···················· | ···················· |
| V | 5866 hCoV-19/Japan/PG-2839/2020\|EPI_ISL_690966\|2020-04 | ···················· | ···················· | ····················· | ···················· | ···················· | ···················· |
| V | 5867 hCoV-19/Brazil/SP-698/2020\|EPI_ISL_693240\|2020-06- | ········T··········· | ···················· | ····················· | ···················· | ···················· | ···················· |
| V | 5868 hCoV-19/Brazil/SP-716/2020\|EPI_ISL_693244\|2020-07- | ···················· | ···················· | ····················· | ···················· | ···················· | ···················· |
| V | 5869 hCoV-19/Brazil/SP-719/2020\|EPI_ISL_693245\|2020-07- | ···················· | ···················· | ····················· | ···················· | ···················· | ···················· |
| V | 5870 hCoV-19/Brazil/SP-721/2020\|EPI_ISL_693246\|2020-06- | ···················· | ···················· | ····················· | ···················· | ···················· | ···················· |
| V | 5871 hCoV-19/Brazil/SP-705/2020\|EPI_ISL_693247\|2020-06- | ···················· | ···················· | ····················· | ···················· | ···················· | ···················· |
| V | 5872 hCoV-19/Brazil/SP-735/2020\|EPI_ISL_693248\|2020-07- | ···················· | ···················· | ····················· | ···················· | ···················· | ···················· |
| V | 5873 hCoV-19/Australia/QLD1245/2020\|EPI_ISL_693250\|2020 | ···················· | ···················· | ····················· | ···················· | ···················· | ···················· |
| V | 5874 hCoV-19/Australia/QLD1246/2020\|EPI_ISL_693251\|2020 | ···················· | ···················· | ····················· | ···················· | ···················· | ···················· |
| V | 5875 hCoV-19/Australia/QLD1248/2020\|EPI_ISL_693252\|2020 | ···················· | ···················· | ····················· | ···················· | ···················· | ···················· |
| V | 5876 hCoV-19/Australia/QLD1249/2020\|EPI_ISL_693253\|2020 | ···················· | ···················· | ····················· | ···················· | ···················· | ···················· |
| V | 5877 hCoV-19/Australia/QLD1251/2020\|EPI_ISL_693254\|2020 | ···················· | ···················· | ····················· | ···················· | ···················· | ···················· |
| V | 5878 hCoV-19/Australia/QLD1252/2020\|EPI_ISL_693255\|2020 | ···················· | ···················· | ····················· | ···················· | ···················· | ···················· |
| V | 5879 hCoV-19/Australia/QLD1253/2020\|EPI_ISL_693256\|2020 | ···················· | ···················· | ····················· | ···················· | ···················· | ···················· |
| V | 5880 hCoV-19/Australia/QLD1254/2020\|EPI_ISL_693257\|2020 | ···················· | ···················· | ····················· | ···················· | ···················· | ···················· |
| V | 5881 hCoV-19/Australia/QLD1255/2020\|EPI_ISL_693258\|2020 | ···················· | ···················· | ····················· | ···················· | ···················· | ···················· |
| V | 5882 hCoV-19/Australia/QLD1256/2020\|EPI_ISL_693259\|2020 | ···················· | ·A·················· | ····················· | ···················· | ···················· | ···················· |
| V | 5883 hCoV-19/Australia/QLD1257/2020\|EPI_ISL_693260\|2020 | ···················· | ···················· | ····················· | ···················· | ···················· | ···················· |
| V | 5884 hCoV-19/Australia/QLD1258/2020\|EPI_ISL_693261\|2020 | ···················· | ···················· | ····················· | ···················· | ···················· | ···················· |
| V | 5885 hCoV-19/Australia/QLD1259/2020\|EPI_ISL_693262\|2020 | ···················· | ···················· | ····················· | ···················· | ···················· | ···················· |
| V | 5886 hCoV-19/Australia/QLD1261/2020\|EPI_ISL_693263\|2020 | ···················· | ···················· | ····················· | ···················· | ···················· | ···················· |
| V | 5887 hCoV-19/Australia/QLD1262/2020\|EPI_ISL_693264\|2020 | ···················· | ···················· | ····················· | ···················· | ···················· | ···················· |
| V | 5888 hCoV-19/Australia/QLD1263/2020\|EPI_ISL_693265\|2020 | ···················· | ···················· | ····················· | ···················· | ···················· | ···················· |
| V | 5889 hCoV-19/Japan/PG-2455/2020\|EPI_ISL_686060\|2020-04 | ···················· | ···················· | ····················· | ···················· | ···················· | ···················· |

| | | | | | | | |
|---|---|---|---|---|---|---|---|
| V | 5890 hCoV-19/Japan/PG-2456/2020\|EPI_ISL_686061\|2020-04 | .................... | .................... | ..................... | .................... | .................... | .................... |
| V | 5891 hCoV-19/Japan/PG-2457/2020\|EPI_ISL_686062\|2020-04 | .................... | .................... | ..................... | .................... | .................... | .................... |
| V | 5892 hCoV-19/Japan/PG-2458/2020\|EPI_ISL_686063\|2020-04 | .................... | .................... | ..................... | .................... | .................... | .................... |
| V | 5893 hCoV-19/Japan/PG-2459/2020\|EPI_ISL_686064\|2020-04 | .................... | .................... | ..................... | .................... | .................... | .................... |
| V | 5894 hCoV-19/Japan/PG-2460/2020\|EPI_ISL_686065\|2020-04 | .................... | .................... | ..................... | .................... | .................... | .................... |
| V | 5895 hCoV-19/Japan/PG-2461/2020\|EPI_ISL_686066\|2020-04 | .................... | .................... | ..................... | .................... | .................... | .................... |
| V | 5896 hCoV-19/Japan/PG-2462/2020\|EPI_ISL_686067\|2020-04 | .................... | .................... | ..................... | .................... | .................... | .................... |
| V | 5897 hCoV-19/Japan/PG-2463/2020\|EPI_ISL_686068\|2020-04 | .................... | .................... | ..................... | .................... | .................... | .................... |
| V | 5898 hCoV-19/Japan/PG-2464/2020\|EPI_ISL_686069\|2020-04 | .................... | .................... | ..................... | .................... | .................... | .................... |
| V | 5899 hCoV-19/Japan/PG-2465/2020\|EPI_ISL_686070\|2020-04 | .................... | .................... | ..................... | .................... | .................... | .................... |
| V | 5900 hCoV-19/Japan/PG-2466/2020\|EPI_ISL_686071\|2020-04 | .................... | .................... | ..................... | .................... | .................... | .................... |
| V | 5901 hCoV-19/Japan/PG-2467/2020\|EPI_ISL_686072\|2020-04 | .................... | .................... | ..................... | .................... | .................... | .................... |
| V | 5902 hCoV-19/Japan/PG-2468/2020\|EPI_ISL_686073\|2020-04 | .................... | .................... | ..................... | .................... | .................... | .................... |
| V | 5903 hCoV-19/Japan/PG-2469/2020\|EPI_ISL_686074\|2020-05 | .................... | .................... | ..................... | .................... | .................... | .................... |
| V | 5904 hCoV-19/Japan/PG-2470/2020\|EPI_ISL_686075\|2020-05 | .................... | .................... | ..................... | .................... | .................... | .................... |
| V | 5905 hCoV-19/Japan/PG-2471/2020\|EPI_ISL_686076\|2020-05 | .................... | .................... | ..................... | .................... | .................... | .................... |
| V | 5906 hCoV-19/Japan/PG-2472/2020\|EPI_ISL_686077\|2020-05 | .................... | .................... | ..................... | .................... | .................... | .................... |
| V | 5907 hCoV-19/Japan/PG-2473/2020\|EPI_ISL_686078\|2020-05 | .................... | .................... | ..................... | .................... | .................... | .................... |
| V | 5908 hCoV-19/Japan/PG-2474/2020\|EPI_ISL_686079\|2020-05 | .................... | .................... | ..................... | .................... | .................... | .................... |
| V | 5909 hCoV-19/Japan/PG-2475/2020\|EPI_ISL_686080\|2020-05 | .................... | .................... | ..................... | .................... | .................... | .................... |
| V | 5910 hCoV-19/Japan/PG-2476/2020\|EPI_ISL_686081\|2020-05 | .................... | .................... | ..................... | .................... | .................... | .................... |
| V | 5911 hCoV-19/Japan/PG-2477/2020\|EPI_ISL_686082\|2020-05 | .................... | .................... | ..................... | .................... | .................... | .................... |
| V | 5912 hCoV-19/Japan/PG-2478/2020\|EPI_ISL_686083\|2020-04 | .................... | .................... | ..................... | .................... | .................... | .................... |
| V | 5913 hCoV-19/Japan/PG-2479/2020\|EPI_ISL_686084\|2020-04 | .................... | .................... | ..................... | .................... | .................... | .................... |
| V | 5914 hCoV-19/Japan/PG-2480/2020\|EPI_ISL_686085\|2020-04 | .................... | .................... | ..................... | .................... | .................... | .................... |
| V | 5915 hCoV-19/Japan/PG-2481/2020\|EPI_ISL_686086\|2020-04 | .................... | .................... | ..................... | .................... | .................... | .................... |
| V | 5916 hCoV-19/Japan/PG-2482/2020\|EPI_ISL_686087\|2020-04 | .................... | .................... | ..................... | .................... | .................... | .................... |
| V | 5917 hCoV-19/Japan/PG-2483/2020\|EPI_ISL_686088\|2020-04 | .................... | .................... | ..................... | .................... | .................... | .................... |
| V | 5918 hCoV-19/Japan/PG-2484/2020\|EPI_ISL_686089\|2020-04 | .................... | .................... | ..................... | .................... | .................... | .................... |
| V | 5919 hCoV-19/Japan/PG-2485/2020\|EPI_ISL_686090\|2020-04 | .................... | .................... | ..................... | .................... | .................... | .................... |
| V | 5920 hCoV-19/Japan/PG-2486/2020\|EPI_ISL_686091\|2020-04 | .................... | .................... | ..................... | .................... | .................... | .................... |
| V | 5921 hCoV-19/Japan/PG-2487/2020\|EPI_ISL_686092\|2020-04 | .................... | .................... | ..................... | .................... | .................... | .................... |
| V | 5922 hCoV-19/Japan/PG-2488/2020\|EPI_ISL_686093\|2020-04 | .................... | .................... | ..................... | .................... | .................... | .................... |
| V | 5923 hCoV-19/Japan/PG-2489/2020\|EPI_ISL_686094\|2020-04 | .................... | .................... | ..................... | .................... | .................... | .................... |
| V | 5924 hCoV-19/Japan/PG-2490/2020\|EPI_ISL_686095\|2020-04 | .................... | .................... | ..................... | .................... | .................... | .................... |
| V | 5925 hCoV-19/Japan/PG-2491/2020\|EPI_ISL_686096\|2020-04 | .................... | .................... | ..................... | .................... | .................... | .................... |
| V | 5926 hCoV-19/Japan/PG-2492/2020\|EPI_ISL_686097\|2020-04 | .................... | .................... | ..................... | .................... | .................... | .................... |
| V | 5927 hCoV-19/Japan/PG-2493/2020\|EPI_ISL_686098\|2020-04 | .................... | .................... | ..................... | .................... | .................... | .................... |
| V | 5928 hCoV-19/Japan/PG-2494/2020\|EPI_ISL_686099\|2020-04 | .................... | .................... | ..................... | .................... | .................... | .................... |
| V | 5929 hCoV-19/Japan/PG-8201/2020\|EPI_ISL_687300\|2020-08 | .................... | .................... | ..................... | .................... | .................... | .................... |
| V | 5930 hCoV-19/Japan/PG-8202/2020\|EPI_ISL_687301\|2020-08 | .................... | .................... | ..................... | .................... | .................... | .................... |
| V | 5931 hCoV-19/Japan/PG-8203/2020\|EPI_ISL_687302\|2020-08 | .................... | .................... | ..................... | .................... | .................... | .................... |
| V | 5932 hCoV-19/Japan/PG-8204/2020\|EPI_ISL_687303\|2020-08 | .................... | .................... | ..................... | .................... | .................... | .................... |
| V | 5933 hCoV-19/Japan/PG-8291/2020\|EPI_ISL_687390\|2020-08 | .................... | .................... | ..................... | .................... | .................... | .................... |
| V | 5934 hCoV-19/Japan/PG-8292/2020\|EPI_ISL_687391\|2020-08 | .................... | .................... | ..................... | .................... | .................... | .................... |
| V | 5935 hCoV-19/Japan/PG-8293/2020\|EPI_ISL_687392\|2020-08 | .................... | .................... | ..................... | .................... | .................... | .................... |
| V | 5936 hCoV-19/Japan/PG-8294/2020\|EPI_ISL_687393\|2020-08 | .................... | .................... | ..................... | .................... | .................... | .................... |
| V | 5937 hCoV-19/Japan/PG-8295/2020\|EPI_ISL_687394\|2020-08 | .................... | .................... | ..................... | .................... | .................... | .................... |
| V | 5938 hCoV-19/Japan/PG-8296/2020\|EPI_ISL_687395\|2020-08 | .................... | .................... | ..................... | .................... | .................... | .................... |
| V | 5939 hCoV-19/Japan/PG-8297/2020\|EPI_ISL_687396\|2020-08 | .................... | .................... | ..................... | .................... | .................... | .................... |
| V | 5940 hCoV-19/Japan/PG-8298/2020\|EPI_ISL_687397\|2020-08 | .................... | .................... | ..................... | .................... | .................... | .................... |
| V | 5941 hCoV-19/Japan/PG-8299/2020\|EPI_ISL_687398\|2020-08 | .................... | .................... | ..................... | .................... | .................... | .................... |
| V | 5942 hCoV-19/Japan/PG-8300/2020\|EPI_ISL_687399\|2020-08 | .................... | .................... | ..................... | .................... | .................... | .................... |
| V | 5943 hCoV-19/Japan/PG-5636/2020\|EPI_ISL_688624\|2020-03 | .................... | .................... | ..................... | .................... | .................... | .................... |
| V | 5944 hCoV-19/Japan/PG-5734/2020\|EPI_ISL_688625\|2020-08 | .................... | .................... | ..................... | .................... | .................... | .................... |
| V | 5945 hCoV-19/Japan/PG-5808/2020\|EPI_ISL_688634\|2020-08 | .................... | .................... | ..................... | .................... | .................... | .................... |
| V | 5946 hCoV-19/Japan/PG-4764/2020\|EPI_ISL_689900\|2020-08 | .................... | .................... | ..................... | .................... | .................... | .................... |
| V | 5947 hCoV-19/Japan/PG-4765/2020\|EPI_ISL_689901\|2020-08 | .................... | .................... | ..................... | .................... | .................... | .................... |
| V | 5948 hCoV-19/Japan/PG-4766/2020\|EPI_ISL_689902\|2020-08 | .................... | .................... | ..................... | .................... | .................... | .................... |
| V | 5949 hCoV-19/Japan/PG-4767/2020\|EPI_ISL_689903\|2020-08 | .................... | .................... | ..................... | .................... | .................... | .................... |
| V | 5950 hCoV-19/Japan/PG-4768/2020\|EPI_ISL_689904\|2020-08 | .................... | .................... | ..................... | .................... | .................... | .................... |
| V | 5951 hCoV-19/Japan/PG-4769/2020\|EPI_ISL_689905\|2020-08 | .................... | .................... | ..................... | .................... | .................... | .................... |
| V | 5952 hCoV-19/Japan/PG-4770/2020\|EPI_ISL_689906\|2020-08 | .................... | .................... | ..................... | .................... | .................... | .................... |
| V | 5953 hCoV-19/Japan/PG-4771/2020\|EPI_ISL_689907\|2020-08 | .................... | .................... | ..................... | .................... | .................... | .................... |
| V | 5954 hCoV-19/Japan/PG-4772/2020\|EPI_ISL_689908\|2020-08 | .................... | .................... | ..................... | .................... | .................... | .................... |
| V | 5955 hCoV-19/Japan/PG-4773/2020\|EPI_ISL_689909\|2020-08 | .................... | .................... | ..................... | .................... | .................... | .................... |
| V | 5956 hCoV-19/Japan/PG-4774/2020\|EPI_ISL_689910\|2020-08 | .................... | .................... | ..................... | .................... | .................... | .................... |
| V | 5957 hCoV-19/Japan/PG-4775/2020\|EPI_ISL_689911\|2020-08 | .................... | .................... | ..................... | .................... | .................... | .................... |
| V | 5958 hCoV-19/Japan/PG-4776/2020\|EPI_ISL_689912\|2020-08 | .................... | .................... | ..................... | .................... | .................... | .................... |
| V | 5959 hCoV-19/Japan/PG-4777/2020\|EPI_ISL_689913\|2020-08 | .................... | .................... | ..................... | .................... | .................... | .................... |
| V | 5960 hCoV-19/Japan/PG-4785/2020\|EPI_ISL_689914\|2020-08 | .................... | .................... | ..................... | .................... | .................... | .................... |
| V | 5961 hCoV-19/Japan/PG-4786/2020\|EPI_ISL_689915\|2020-08 | .................... | .................... | ..................... | .................... | .................... | .................... |
| V | 5962 hCoV-19/Japan/PG-4787/2020\|EPI_ISL_689916\|2020-08 | .................... | .................... | ..................... | .................... | .................... | .................... |
| V | 5963 hCoV-19/Japan/PG-4788/2020\|EPI_ISL_689917\|2020-08 | .................... | .................... | ..................... | .................... | .................... | .................... |
| V | 5964 hCoV-19/Japan/PG-4793/2020\|EPI_ISL_689918\|2020-08 | .................... | .................... | ..................... | .................... | .................... | .................... |
| V | 5965 hCoV-19/Japan/PG-4794/2020\|EPI_ISL_689919\|2020-08 | .................... | .................... | ..................... | .................... | .................... | .................... |
| V | 5966 hCoV-19/Japan/PG-4795/2020\|EPI_ISL_689920\|2020-08 | .................... | .................... | ..................... | .................... | .................... | .................... |
| V | 5967 hCoV-19/Japan/PG-4796/2020\|EPI_ISL_689921\|2020-08 | .................... | .................... | ..................... | .................... | .................... | .................... |
| V | 5968 hCoV-19/Japan/PG-4797/2020\|EPI_ISL_689922\|2020-08 | .................... | .................... | ..................... | .................... | .................... | .................... |
| V | 5969 hCoV-19/Japan/PG-4798/2020\|EPI_ISL_689923\|2020-08 | .................... | .................... | ..................... | .................... | .................... | .................... |

| | | | | | | | | |
|---|---|---|---|---|---|---|---|---|
| V | 5970 | hCoV-19/Japan/PG-4799/2020\|EPI_ISL_689924\|2020-08 | .................... | .................... | ..................... | .................... | .................... | .................... |
| V | 5971 | hCoV-19/Japan/PG-4800/2020\|EPI_ISL_689925\|2020-08 | .................... | .................... | ..................... | .................... | .................... | .................... |
| V | 5972 | hCoV-19/Japan/PG-4801/2020\|EPI_ISL_689926\|2020-07 | .................... | .................... | ..................... | .................... | .................... | .................... |
| V | 5973 | hCoV-19/Japan/PG-4802/2020\|EPI_ISL_689927\|2020-08 | .................... | .................... | ..................... | .................... | .................... | .................... |
| V | 5974 | hCoV-19/Japan/PG-4803/2020\|EPI_ISL_689928\|2020-08 | .................... | .................... | ..................... | .................... | .................... | .................... |
| V | 5975 | hCoV-19/Japan/PG-4804/2020\|EPI_ISL_689929\|2020-08 | .................... | .................... | ..................... | .................... | .................... | .................... |
| V | 5976 | hCoV-19/Japan/PG-4806/2020\|EPI_ISL_689930\|2020-08 | .................... | .................... | ..................... | .................... | .................... | .................... |
| V | 5977 | hCoV-19/Japan/PG-4807/2020\|EPI_ISL_689931\|2020-08 | .................... | .................... | ..................... | .................... | .................... | .................... |
| V | 5978 | hCoV-19/Japan/PG-4808/2020\|EPI_ISL_689932\|2020-08 | .................... | .................... | ..................... | .................... | .................... | .................... |
| V | 5979 | hCoV-19/Japan/PG-4809/2020\|EPI_ISL_689933\|2020-08 | .................... | .................... | ..................... | .................... | .................... | .................... |
| V | 5980 | hCoV-19/Japan/PG-4810/2020\|EPI_ISL_689934\|2020-08 | .................... | .................... | ..................... | .................... | .................... | .................... |
| V | 5981 | hCoV-19/Japan/PG-4811/2020\|EPI_ISL_689935\|2020-08 | .................... | .................... | ..................... | .................... | .................... | .................... |
| V | 5982 | hCoV-19/Japan/PG-4812/2020\|EPI_ISL_689936\|2020-08 | .................... | .................... | ..................... | .................... | .................... | .................... |
| V | 5983 | hCoV-19/Japan/PG-4813/2020\|EPI_ISL_689937\|2020-08 | .................... | .................... | ..................... | .................... | .................... | .................... |
| V | 5984 | hCoV-19/Japan/PG-4814/2020\|EPI_ISL_689938\|2020-08 | .................... | .................... | ..................... | .................... | .................... | .................... |
| V | 5985 | hCoV-19/Japan/PG-4815/2020\|EPI_ISL_689939\|2020-08 | .................... | .................... | ..................... | .................... | .................... | .................... |
| V | 5986 | hCoV-19/Japan/PG-4816/2020\|EPI_ISL_689940\|2020-08 | .................... | .................... | ..................... | .................... | .................... | .................... |
| V | 5987 | hCoV-19/Japan/PG-4817/2020\|EPI_ISL_689941\|2020-08 | .................... | .................... | ..................... | .................... | .................... | .................... |
| V | 5988 | hCoV-19/Japan/PG-4818/2020\|EPI_ISL_689942\|2020-07 | .................... | .................... | ..................... | .................... | .................... | .................... |
| V | 5989 | hCoV-19/Japan/PG-4819/2020\|EPI_ISL_689943\|2020-07 | .................... | .................... | ..................... | .................... | .................... | .................... |
| V | 5990 | hCoV-19/Japan/PG-4820/2020\|EPI_ISL_689944\|2020-07 | .................... | .................... | ..................... | .................... | .................... | .................... |
| V | 5991 | hCoV-19/Japan/PG-4821/2020\|EPI_ISL_689945\|2020-07 | .................... | .................... | ..................... | .................... | .................... | .................... |
| V | 5992 | hCoV-19/Japan/PG-4822/2020\|EPI_ISL_689946\|2020-07 | .................... | .................... | ..................... | .................... | .................... | .................... |
| V | 5993 | hCoV-19/Japan/PG-4823/2020\|EPI_ISL_689947\|2020-07 | .................... | .................... | ..................... | .................... | .................... | .................... |
| V | 5994 | hCoV-19/Japan/PG-4824/2020\|EPI_ISL_689948\|2020-07 | .................... | .................... | ..................... | .................... | .................... | .................... |
| V | 5995 | hCoV-19/Japan/PG-4825/2020\|EPI_ISL_689949\|2020-07 | .................... | .................... | ..................... | .................... | .................... | .................... |
| V | 5996 | hCoV-19/Japan/PG-4826/2020\|EPI_ISL_689950\|2020-07 | .................... | .................... | ..................... | .................... | .................... | .................... |
| V | 5997 | hCoV-19/Japan/PG-4827/2020\|EPI_ISL_689951\|2020-07 | .................... | .................... | ..................... | .................... | .................... | .................... |
| V | 5998 | hCoV-19/Japan/PG-4828/2020\|EPI_ISL_689952\|2020-07 | .................... | .................... | ..................... | .................... | .................... | .................... |
| V | 5999 | hCoV-19/Japan/PG-4829/2020\|EPI_ISL_689953\|2020-07 | .................... | .................... | ..................... | .................... | .................... | .................... |
| V | 6000 | hCoV-19/Japan/PG-4830/2020\|EPI_ISL_689954\|2020-07 | .................... | .................... | ..................... | .................... | .................... | .................... |
| V | 6001 | hCoV-19/Japan/PG-4831/2020\|EPI_ISL_689955\|2020-07 | .................... | .................... | ..................... | .................... | .................... | .................... |
| V | 6002 | hCoV-19/Japan/PG-4832/2020\|EPI_ISL_689956\|2020-07 | .................... | .................... | ..................... | .................... | .................... | .................... |
| V | 6003 | hCoV-19/Japan/PG-4833/2020\|EPI_ISL_689957\|2020-07 | .................... | .................... | ..................... | .................... | .................... | .................... |
| V | 6004 | hCoV-19/Japan/PG-4834/2020\|EPI_ISL_689958\|2020-07 | .................... | .................... | ..................... | .................... | .................... | .................... |
| V | 6005 | hCoV-19/Japan/PG-4835/2020\|EPI_ISL_689959\|2020-07 | .................... | .................... | ..................... | .................... | .................... | .................... |
| V | 6006 | hCoV-19/Japan/PG-4836/2020\|EPI_ISL_689960\|2020-07 | .................... | .................... | ..................... | .................... | .................... | .................... |
| V | 6007 | hCoV-19/Japan/PG-4839/2020\|EPI_ISL_689961\|2020-07 | .................... | .................... | ..................... | .................... | .................... | .................... |
| V | 6008 | hCoV-19/Japan/PG-4840/2020\|EPI_ISL_689962\|2020-07 | .................... | .................... | ..................... | .................... | .................... | .................... |
| V | 6009 | hCoV-19/Japan/PG-4841/2020\|EPI_ISL_689963\|2020-08 | .................... | .................... | ..................... | .................... | .................... | .................... |
| V | 6010 | hCoV-19/Japan/PG-4842/2020\|EPI_ISL_689964\|2020-08 | .................... | .................... | ..................... | .................... | .................... | .................... |
| V | 6011 | hCoV-19/Japan/PG-4843/2020\|EPI_ISL_689965\|2020-07 | .................... | .................... | ..................... | .................... | .................... | .................... |
| V | 6012 | hCoV-19/Japan/PG-4856/2020\|EPI_ISL_689978\|2020-07 | .................... | .................... | ..................... | .................... | .................... | .................... |
| V | 6013 | hCoV-19/Japan/PG-4858/2020\|EPI_ISL_689979\|2020-08 | .................... | .................... | ..................... | .................... | .................... | .................... |
| V | 6014 | hCoV-19/Japan/PG-4859/2020\|EPI_ISL_689980\|2020-08 | .................... | .................... | ..................... | .................... | .................... | .................... |
| V | 6015 | hCoV-19/Japan/PG-4860/2020\|EPI_ISL_689981\|2020-08 | .................... | .................... | ..................... | .................... | .................... | .................... |
| V | 6016 | hCoV-19/Japan/PG-4861/2020\|EPI_ISL_689982\|2020-08 | .................... | .................... | ..................... | .................... | .................... | .................... |
| V | 6017 | hCoV-19/Japan/PG-4862/2020\|EPI_ISL_689983\|2020-08 | .................... | .................... | ..................... | .................... | .................... | .................... |
| V | 6018 | hCoV-19/Japan/PG-4863/2020\|EPI_ISL_689984\|2020-07 | .................... | .................... | ..................... | .................... | .................... | .................... |
| V | 6019 | hCoV-19/Japan/PG-4864/2020\|EPI_ISL_689985\|2020-07 | .................... | .................... | ..................... | .................... | .................... | .................... |
| V | 6020 | hCoV-19/Japan/PG-4865/2020\|EPI_ISL_689986\|2020-07 | .................... | .................... | ..................... | .................... | .................... | .................... |
| V | 6021 | hCoV-19/Japan/PG-4866/2020\|EPI_ISL_689987\|2020-07 | .................... | .................... | ..................... | .................... | .................... | .................... |
| V | 6022 | hCoV-19/Japan/PG-4867/2020\|EPI_ISL_689988\|2020-08 | .................... | .................... | ..................... | .................... | .................... | .................... |
| V | 6023 | hCoV-19/Japan/PG-4868/2020\|EPI_ISL_689989\|2020-08 | .................... | .................... | ..................... | .................... | .................... | .................... |
| V | 6024 | hCoV-19/Japan/PG-4869/2020\|EPI_ISL_689990\|2020-08 | .................... | .................... | ..................... | .................... | .................... | .................... |
| V | 6025 | hCoV-19/Japan/PG-4870/2020\|EPI_ISL_689991\|2020-07 | .................... | .................... | ..................... | .................... | .................... | .................... |
| V | 6026 | hCoV-19/Japan/PG-4871/2020\|EPI_ISL_689992\|2020-08 | .................... | .................... | ..................... | .................... | .................... | .................... |
| V | 6027 | hCoV-19/Japan/PG-4872/2020\|EPI_ISL_689993\|2020-08 | .................... | .................... | ..................... | .................... | .................... | .................... |
| V | 6028 | hCoV-19/Japan/PG-4873/2020\|EPI_ISL_689994\|2020-08 | .................... | .................... | ..................... | .................... | .................... | .................... |
| V | 6029 | hCoV-19/Japan/PG-4874/2020\|EPI_ISL_689995\|2020-08 | .................... | .................... | ..................... | .................... | .................... | .................... |
| V | 6030 | hCoV-19/Japan/PG-4875/2020\|EPI_ISL_689996\|2020-08 | .................... | .................... | ..................... | .................... | .................... | .................... |
| V | 6031 | hCoV-19/Japan/PG-4876/2020\|EPI_ISL_689997\|2020-08 | .................... | .................... | ..................... | .................... | .................... | .................... |
| V | 6032 | hCoV-19/Japan/PG-4877/2020\|EPI_ISL_689998\|2020-08 | .................... | .................... | ..................... | .................... | .................... | .................... |
| V | 6033 | hCoV-19/Japan/PG-4878/2020\|EPI_ISL_689999\|2020-08 | .................... | .................... | ..................... | .................... | .................... | .................... |
| V | 6034 | hCoV-19/Japan/PG-2768/2020\|EPI_ISL_690900\|2020-04 | .................... | .................... | ..................... | .................... | .................... | .................... |
| V | 6035 | hCoV-19/Japan/PG-2769/2020\|EPI_ISL_690901\|2020-04 | .................... | .................... | ..................... | .................... | .................... | .................... |
| V | 6036 | hCoV-19/Japan/PG-2775/2020\|EPI_ISL_690902\|2020-06 | .................... | .................... | ..................... | .................... | .................... | .................... |
| V | 6037 | hCoV-19/Japan/PG-2776/2020\|EPI_ISL_690903\|2020-06 | .................... | .................... | ..................... | .................... | .................... | .................... |
| V | 6038 | hCoV-19/Japan/PG-2777/2020\|EPI_ISL_690904\|2020-06 | .................... | .................... | ..................... | .................... | .................... | .................... |
| V | 6039 | hCoV-19/Japan/PG-2778/2020\|EPI_ISL_690905\|2020-06 | .................... | .................... | ..................... | .................... | .................... | .................... |
| V | 6040 | hCoV-19/Japan/PG-2779/2020\|EPI_ISL_690906\|2020-06 | .................... | .................... | ..................... | .................... | .................... | .................... |
| V | 6041 | hCoV-19/Japan/PG-2780/2020\|EPI_ISL_690907\|2020-06 | .................... | .................... | ..................... | .................... | .................... | .................... |
| V | 6042 | hCoV-19/Japan/PG-2781/2020\|EPI_ISL_690908\|2020-06 | .................... | .................... | ..................... | .................... | .................... | .................... |
| V | 6043 | hCoV-19/Japan/PG-2782/2020\|EPI_ISL_690909\|2020-04 | .................... | .................... | ..................... | .................... | .................... | .................... |
| V | 6044 | hCoV-19/Japan/PG-2783/2020\|EPI_ISL_690910\|2020-04 | .................... | .................... | ..................... | .................... | .................... | .................... |
| V | 6045 | hCoV-19/Japan/PG-2784/2020\|EPI_ISL_690911\|2020-04 | .................... | .................... | ..................... | .................... | .................... | .................... |
| V | 6046 | hCoV-19/Japan/PG-2785/2020\|EPI_ISL_690912\|2020-04 | .................... | .A.................. | ..................... | .................... | .................... | .................... |
| V | 6047 | hCoV-19/Japan/PG-2786/2020\|EPI_ISL_690913\|2020-04 | .................... | .A.................. | ..................... | .................... | .................... | .................... |
| V | 6048 | hCoV-19/Japan/PG-2787/2020\|EPI_ISL_690914\|2020-04 | .................... | .................... | ..................... | .................... | .................... | .................... |
| V | 6049 | hCoV-19/Japan/PG-2788/2020\|EPI_ISL_690915\|2020-04 | .................... | .................... | ..................... | .................... | .................... | .................... |

| | | | | | | | | |
|---|---|---|---|---|---|---|---|---|
| V | 6050 | hCoV-19/Japan/PG-2789/2020\|EPI_ISL_690916\|2020-04 | ···················· | ···················· | ····················· | ···················· | ···················· | ···················· |
| V | 6051 | hCoV-19/Japan/PG-2790/2020\|EPI_ISL_690917\|2020-04 | ···················· | ···················· | ····················· | ···················· | ···················· | ···················· |
| V | 6052 | hCoV-19/Japan/PG-2791/2020\|EPI_ISL_690918\|2020-04 | ···················· | ···················· | ····················· | ···················· | ···················· | ···················· |
| V | 6053 | hCoV-19/Japan/PG-2792/2020\|EPI_ISL_690919\|2020-04 | ···················· | ·A·················· | ····················· | ···················· | ···················· | ···················· |
| V | 6054 | hCoV-19/Japan/PG-2793/2020\|EPI_ISL_690920\|2020-04 | ···················· | ···················· | ····················· | ···················· | ···················· | ···················· |
| V | 6055 | hCoV-19/Japan/PG-2794/2020\|EPI_ISL_690921\|2020-04 | ···················· | ·A·················· | ····················· | ···················· | ···················· | ···················· |
| V | 6056 | hCoV-19/Japan/PG-2795/2020\|EPI_ISL_690922\|2020-04 | ···················· | ···················· | ····················· | ···················· | ···················· | ···················· |
| V | 6057 | hCoV-19/Japan/PG-2796/2020\|EPI_ISL_690923\|2020-04 | ···················· | ···················· | ····················· | ···················· | ···················· | ···················· |
| V | 6058 | hCoV-19/Japan/PG-2797/2020\|EPI_ISL_690924\|2020-04 | ···················· | ···················· | ····················· | ···················· | ···················· | ···················· |
| V | 6059 | hCoV-19/Japan/PG-2798/2020\|EPI_ISL_690925\|2020-04 | ···················· | ···················· | ····················· | ···················· | ···················· | ···················· |
| V | 6060 | hCoV-19/Japan/PG-2799/2020\|EPI_ISL_690926\|2020-04 | ···················· | ···················· | ····················· | ···················· | ···················· | ···················· |
| V | 6061 | hCoV-19/Japan/PG-2800/2020\|EPI_ISL_690927\|2020-04 | ···················· | ···················· | ····················· | ···················· | ···················· | ···················· |
| V | 6062 | hCoV-19/Japan/PG-2801/2020\|EPI_ISL_690928\|2020-04 | ···················· | ···················· | ····················· | ···················· | ···················· | ···················· |
| V | 6063 | hCoV-19/Japan/PG-2802/2020\|EPI_ISL_690929\|2020-04 | ···················· | ···················· | ····················· | ···················· | ···················· | ···················· |
| V | 6064 | hCoV-19/Japan/PG-2803/2020\|EPI_ISL_690930\|2020-04 | ···················· | ···················· | ····················· | ···················· | ···················· | ···················· |
| V | 6065 | hCoV-19/Japan/PG-2804/2020\|EPI_ISL_690931\|2020-04 | ···················· | ···················· | ····················· | ···················· | ···················· | ···················· |
| V | 6066 | hCoV-19/Japan/PG-2805/2020\|EPI_ISL_690932\|2020-04 | ···················· | ···················· | ····················· | ···················· | ···················· | ···················· |
| V | 6067 | hCoV-19/Japan/PG-2806/2020\|EPI_ISL_690933\|2020-04 | ···················· | ···················· | ····················· | ···················· | ···················· | ···················· |
| V | 6068 | hCoV-19/Japan/PG-2807/2020\|EPI_ISL_690934\|2020-04 | ···················· | ···················· | ····················· | ···················· | ···················· | ···················· |
| V | 6069 | hCoV-19/Japan/PG-2808/2020\|EPI_ISL_690935\|2020-04 | ···················· | ···················· | ····················· | ···················· | ···················· | ···················· |
| V | 6070 | hCoV-19/Japan/PG-2809/2020\|EPI_ISL_690936\|2020-04 | ···················· | ···················· | ····················· | ···················· | ···················· | ···················· |
| V | 6072 | hCoV-19/Japan/PG-2811/2020\|EPI_ISL_690938\|2020-04 | ···················· | ···················· | ····················· | ···················· | ···················· | ···················· |
| V | 6073 | hCoV-19/Japan/PG-2812/2020\|EPI_ISL_690939\|2020-04 | ···················· | ···················· | ····················· | ···················· | ···················· | ···················· |
| V | 6074 | hCoV-19/Japan/PG-2813/2020\|EPI_ISL_690940\|2020-04 | ···················· | ···················· | ····················· | ···················· | ···················· | ···················· |
| V | 6075 | hCoV-19/Japan/PG-2814/2020\|EPI_ISL_690941\|2020-04 | ···················· | ···················· | ····················· | ···················· | ···················· | ···················· |
| V | 6076 | hCoV-19/Japan/PG-2815/2020\|EPI_ISL_690942\|2020-04 | ···················· | ···················· | ····················· | ···················· | ···················· | ···················· |
| V | 6077 | hCoV-19/Japan/PG-2816/2020\|EPI_ISL_690943\|2020-04 | ···················· | ···················· | ····················· | ···················· | ···················· | ···················· |
| V | 6078 | hCoV-19/Japan/PG-2817/2020\|EPI_ISL_690944\|2020-04 | ···················· | ···················· | ····················· | ···················· | ···················· | ···················· |
| V | 6079 | hCoV-19/Japan/PG-2818/2020\|EPI_ISL_690945\|2020-04 | ···················· | ···················· | ····················· | ···················· | ···················· | ···················· |
| V | 6080 | hCoV-19/Japan/PG-2819/2020\|EPI_ISL_690946\|2020-04 | ···················· | ···················· | ····················· | ···················· | ···················· | ···················· |
| V | 6081 | hCoV-19/Japan/PG-2820/2020\|EPI_ISL_690947\|2020-04 | ···················· | ···················· | ····················· | ···················· | ···················· | ···················· |
| V | 6082 | hCoV-19/Japan/PG-2821/2020\|EPI_ISL_690948\|2020-04 | ···················· | ···················· | ····················· | ···················· | ···················· | ···················· |
| V | 6083 | hCoV-19/Japan/PG-2822/2020\|EPI_ISL_690949\|2020-04 | ···················· | ···················· | ····················· | ···················· | ···················· | ···················· |
| V | 6084 | hCoV-19/Japan/PG-2833/2020\|EPI_ISL_690960\|2020-04 | ···················· | ···················· | ····················· | ···················· | ···················· | ···················· |
| V | 6085 | hCoV-19/Japan/PG-2834/2020\|EPI_ISL_690961\|2020-04 | ···················· | ·A·················· | ····················· | ···················· | ···················· | ···················· |
| V | 6086 | hCoV-19/Japan/PG-2835/2020\|EPI_ISL_690962\|2020-04 | ···················· | ·A·················· | ····················· | ···················· | ···················· | ···················· |
| V | 6087 | hCoV-19/Japan/PG-2836/2020\|EPI_ISL_690963\|2020-04 | ···················· | ···················· | ····················· | ···················· | ···················· | ···················· |
| V | 6088 | hCoV-19/Japan/PG-2837/2020\|EPI_ISL_690964\|2020-04 | ···················· | ···················· | ····················· | ···················· | ···················· | ···················· |
| V | 6089 | hCoV-19/Japan/PG-2838/2020\|EPI_ISL_690965\|2020-04 | ···················· | ·A·················· | ····················· | ···················· | ···················· | ···················· |
| V | 6090 | hCoV-19/Japan/PG-2840/2020\|EPI_ISL_690967\|2020-04 | ···················· | ···················· | ····················· | ···················· | ···················· | ···················· |
| V | 6091 | hCoV-19/Japan/PG-2841/2020\|EPI_ISL_690968\|2020-04 | ···················· | ···················· | ····················· | ···················· | ···················· | ···················· |
| V | 6092 | hCoV-19/Japan/PG-2842/2020\|EPI_ISL_690969\|2020-04 | ···················· | ···················· | ····················· | ···················· | ···················· | ···················· |
| V | 6093 | hCoV-19/Japan/PG-2843/2020\|EPI_ISL_690970\|2020-04 | ···················· | ·A·················· | ····················· | ···················· | ···················· | ···················· |
| V | 6094 | hCoV-19/Japan/PG-2844/2020\|EPI_ISL_690971\|2020-04 | ···················· | ···················· | ····················· | ···················· | ···················· | ···················· |
| V | 6095 | hCoV-19/Japan/PG-2845/2020\|EPI_ISL_690972\|2020-04 | ···················· | ···················· | ····················· | ···················· | ···················· | ···················· |
| V | 6096 | hCoV-19/Japan/PG-2846/2020\|EPI_ISL_690973\|2020-05 | ···················· | ···················· | ····················· | ···················· | ···················· | ···················· |
| V | 6097 | hCoV-19/Japan/PG-2847/2020\|EPI_ISL_690974\|2020-05 | ···················· | ·A·················· | ····················· | ···················· | ···················· | ···················· |
| V | 6098 | hCoV-19/Japan/PG-2849/2020\|EPI_ISL_690975\|2020-05 | ···················· | ·A·················· | ····················· | ···················· | ···················· | ···················· |
| V | 6099 | hCoV-19/Japan/PG-2850/2020\|EPI_ISL_690976\|2020-05 | ···················· | ···················· | ····················· | ···················· | ···················· | ···················· |
| V | 6100 | hCoV-19/Japan/PG-2851/2020\|EPI_ISL_690977\|2020-05 | ···················· | ·A·················· | ····················· | ···················· | ···················· | ···················· |
| V | 6101 | hCoV-19/Japan/PG-2852/2020\|EPI_ISL_690978\|2020-04 | ···················· | ···················· | ····················· | ···················· | ···················· | ···················· |
| V | 6102 | hCoV-19/Japan/PG-2853/2020\|EPI_ISL_690979\|2020-04 | ···················· | ···················· | ····················· | ···················· | ···················· | ···················· |
| V | 6103 | hCoV-19/Japan/PG-2854/2020\|EPI_ISL_690980\|2020-04 | ···················· | ···················· | ····················· | ···················· | ···················· | ···················· |
| V | 6104 | hCoV-19/Japan/PG-2855/2020\|EPI_ISL_690981\|2020-06 | ···················· | ···················· | ····················· | ···················· | ···················· | ···················· |
| V | 6105 | hCoV-19/Japan/PG-2856/2020\|EPI_ISL_690982\|2020-04 | ···················· | ···················· | ····················· | ···················· | ···················· | ···················· |
| V | 6106 | hCoV-19/Japan/PG-2857/2020\|EPI_ISL_690983\|2020-04 | ···················· | ···················· | ····················· | ···················· | ···················· | ···················· |
| V | 6107 | hCoV-19/Japan/PG-2858/2020\|EPI_ISL_690984\|2020-04 | ···················· | ···················· | ····················· | ···················· | ···················· | ···················· |
| V | 6108 | hCoV-19/Japan/PG-2859/2020\|EPI_ISL_690985\|2020-04 | ···················· | ···················· | ····················· | ···················· | ···················· | ···················· |
| V | 6109 | hCoV-19/Japan/PG-2860/2020\|EPI_ISL_690986\|2020-04 | ···················· | ···················· | ····················· | ···················· | ···················· | ···················· |
| V | 6110 | hCoV-19/Japan/PG-2861/2020\|EPI_ISL_690987\|2020-03 | ···················· | ···················· | ····················· | ···················· | ···················· | ···················· |
| V | 6111 | hCoV-19/Japan/PG-2862/2020\|EPI_ISL_690988\|2020-03 | ···················· | ···················· | ····················· | ···················· | ···················· | ···················· |
| V | 6112 | hCoV-19/Japan/PG-2863/2020\|EPI_ISL_690989\|2020-03 | ···················· | ·A·················· | ····················· | ···················· | ···················· | ···················· |
| V | 6113 | hCoV-19/Japan/PG-2864/2020\|EPI_ISL_690990\|2020-03 | ···················· | ···················· | ····················· | ···················· | ···················· | ···················· |
| V | 6114 | hCoV-19/Japan/PG-2865/2020\|EPI_ISL_690991\|2020-04 | ···················· | ···················· | ····················· | ···················· | ···················· | ···················· |
| V | 6115 | hCoV-19/Japan/PG-2866/2020\|EPI_ISL_690992\|2020-04 | ···················· | ···················· | ····················· | ···················· | ···················· | ···················· |
| V | 6116 | hCoV-19/Japan/PG-2867/2020\|EPI_ISL_690993\|2020-04 | ···················· | ···················· | ····················· | ···················· | ···················· | ···················· |
| V | 6117 | hCoV-19/Japan/PG-2868/2020\|EPI_ISL_690994\|2020-04 | ···················· | ···················· | ····················· | ···················· | ···················· | ···················· |
| V | 6118 | hCoV-19/Japan/PG-2869/2020\|EPI_ISL_690995\|2020-04 | ···················· | ···················· | ····················· | ···················· | ···················· | ···················· |
| V | 6119 | hCoV-19/Japan/PG-2870/2020\|EPI_ISL_690996\|2020-04 | ···················· | ···················· | ····················· | ···················· | ···················· | ···················· |
| V | 6120 | hCoV-19/Japan/PG-2871/2020\|EPI_ISL_690997\|2020-04 | ···················· | ···················· | ····················· | ···················· | ···················· | ···················· |
| V | 6121 | hCoV-19/Japan/PG-2872/2020\|EPI_ISL_690998\|2020-04 | ···················· | ···················· | ····················· | ···················· | ···················· | ···················· |
| V | 6123 | hCoV-19/USA/MA-MASPHL-00401/2020\|EPI_ISL_693145\|20 | ···················· | ···················· | ····················· | ···················· | ···················· | ···················· |
| V | 6124 | hCoV-19/USA/MA-MASPHL-00400/2020\|EPI_ISL_693146\|20 | ···················· | ···················· | ····················· | ···················· | ···················· | ···················· |
| V | 6125 | hCoV-19/USA/MA-MASPHL-00399/2020\|EPI_ISL_693147\|20 | ···················· | ···················· | ····················· | ···················· | ···················· | ···················· |
| V | 6126 | hCoV-19/USA/MA-MASPHL-00393/2020\|EPI_ISL_693152\|20 | ···················· | ···················· | ····················· | ···················· | ···················· | ···················· |
| V | 6127 | hCoV-19/USA/MA-MASPHL-00386/2020\|EPI_ISL_693158\|20 | ···················· | ···················· | ····················· | ···················· | ···················· | ···················· |
| V | 6128 | hCoV-19/USA/MA-MASPHL-00374/2020\|EPI_ISL_693162\|20 | ···················· | ···················· | ····················· | ···················· | ···················· | ···················· |
| V | 6129 | hCoV-19/USA/MA-MASPHL-00366/2020\|EPI_ISL_693167\|20 | ···················· | ···················· | ····················· | ···················· | ···················· | ···················· |
| V | 6130 | hCoV-19/USA/MA-MASPHL-00356/2020\|EPI_ISL_693176\|20 | ···················· | ···················· | ····················· | ···················· | ···················· | ···················· |
| V | 6131 | hCoV-19/USA/MA-MASPHL-00355/2020\|EPI_ISL_693177\|20 | ···················· | ···················· | ····················· | ···················· | ···················· | ···················· |

| | | | | | | | | |
|---|---|---|---|---|---|---|---|---|
| V | 6132 | hCoV-19/USA/MA-MASPHL-00354/2020\|EPI_ISL_693178\|20 | ···················· | ···················· | ···················· | ···················· | ···················· | ···················· |
| V | 6133 | hCoV-19/USA/MA-MASPHL-00353/2020\|EPI_ISL_693179\|20 | ···················· | ···················· | ···················· | ···················· | ···················· | ···················· |
| V | 6134 | hCoV-19/USA/MA-MASPHL-00350/2020\|EPI_ISL_693182\|20 | ···················· | ···················· | ···················· | ···················· | ···················· | ···················· |
| V | 6135 | hCoV-19/USA/MA-MASPHL-00329/2020\|EPI_ISL_693188\|20 | ···················· | ···················· | ···················· | ···················· | ···················· | ···················· |
| V | 6136 | hCoV-19/USA/MA-MASPHL-00328/2020\|EPI_ISL_693189\|20 | ···················· | ···················· | ···················· | ···················· | ···················· | ···················· |
| V | 6137 | hCoV-19/USA/MA-MASPHL-00327/2020\|EPI_ISL_693190\|20 | ···················· | ···················· | ···················· | ···················· | ···················· | ···················· |
| V | 6138 | hCoV-19/Australia/TAS000280/2020\|EPI_ISL_693192\|20 | ···················· | ···················· | ···················· | ···················· | ···················· | ···················· |
| V | 6139 | hCoV-19/Australia/TAS000281/2020\|EPI_ISL_693193\|20 | ···················· | ···················· | ···················· | ···················· | ···················· | ···················· |
| V | 6140 | hCoV-19/Australia/TAS000282/2020\|EPI_ISL_693194\|20 | ···················· | ···················· | ···················· | ···················· | ···················· | ···················· |
| V | 6141 | hCoV-19/Brazil/SP-565/2020\|EPI_ISL_693195\|2020-03- | ···················· | ···················· | ···················· | ···················· | ···················· | ···················· |
| V | 6142 | hCoV-19/Brazil/SP-243/2020\|EPI_ISL_693196\|2020-05- | ···················· | ···················· | ···················· | ···················· | ·········T·········· | ···················· |
| V | 6143 | hCoV-19/Brazil/SP-249/2020\|EPI_ISL_693198\|2020-05- | ···················· | ···················· | ···················· | ···················· | ···················· | ···················· |
| V | 6144 | hCoV-19/Brazil/SP-253/2020\|EPI_ISL_693199\|2020-05- | ···················· | ···················· | ···················· | ···················· | ···················· | ···················· |
| V | 6145 | hCoV-19/Japan/PG-8101/2020\|EPI_ISL_687200\|2020-08 | ···················· | ···················· | ···················· | ···················· | ···················· | ···················· |
| V | 6146 | hCoV-19/Japan/PG-8102/2020\|EPI_ISL_687201\|2020-08 | ···················· | ···················· | ···················· | ···················· | ···················· | ···················· |
| V | 6147 | hCoV-19/Japan/PG-8103/2020\|EPI_ISL_687202\|2020-08 | ···················· | ···················· | ···················· | ···················· | ···················· | ···················· |
| V | 6148 | hCoV-19/Japan/PG-8104/2020\|EPI_ISL_687203\|2020-08 | ···················· | ···················· | ···················· | ···················· | ···················· | ···················· |
| V | 6149 | hCoV-19/Japan/PG-8105/2020\|EPI_ISL_687204\|2020-08 | ···················· | ···················· | ···················· | ···················· | ···················· | ···················· |
| V | 6150 | hCoV-19/Japan/PG-8106/2020\|EPI_ISL_687205\|2020-08 | ···················· | ···················· | ···················· | ···················· | ···················· | ···················· |
| V | 6151 | hCoV-19/Japan/PG-8107/2020\|EPI_ISL_687206\|2020-08 | ···················· | ···················· | ···················· | ···················· | ···················· | ···················· |
| V | 6152 | hCoV-19/Japan/PG-8108/2020\|EPI_ISL_687207\|2020-08 | ···················· | ···················· | ···················· | ···················· | ···················· | ···················· |
| V | 6153 | hCoV-19/Japan/PG-8109/2020\|EPI_ISL_687208\|2020-08 | ···················· | ···················· | ···················· | ···················· | ···················· | ···················· |
| V | 6154 | hCoV-19/Japan/PG-8110/2020\|EPI_ISL_687209\|2020-08 | ···················· | ···················· | ···················· | ···················· | ···················· | ···················· |
| V | 6155 | hCoV-19/Japan/PG-8111/2020\|EPI_ISL_687210\|2020-08 | ···················· | ···················· | ···················· | ···················· | ···················· | ···················· |
| V | 6156 | hCoV-19/Japan/PG-8112/2020\|EPI_ISL_687211\|2020-08 | ···················· | ···················· | ···················· | ···················· | ···················· | ···················· |
| V | 6157 | hCoV-19/Japan/PG-8113/2020\|EPI_ISL_687212\|2020-08 | ···················· | ···················· | ···················· | ···················· | ···················· | ···················· |
| V | 6158 | hCoV-19/Japan/PG-8114/2020\|EPI_ISL_687213\|2020-08 | ···················· | ···················· | ···················· | ···················· | ···················· | ···················· |
| V | 6159 | hCoV-19/Japan/PG-8115/2020\|EPI_ISL_687214\|2020-08 | ···················· | ···················· | ···················· | ···················· | ···················· | ···················· |
| V | 6160 | hCoV-19/Japan/PG-8116/2020\|EPI_ISL_687215\|2020-08 | ···················· | ···················· | ···················· | ···················· | ···················· | ···················· |
| V | 6161 | hCoV-19/Japan/PG-8117/2020\|EPI_ISL_687216\|2020-08 | ···················· | ···················· | ···················· | ···················· | ···················· | ···················· |
| V | 6162 | hCoV-19/Japan/PG-8118/2020\|EPI_ISL_687217\|2020-08 | ···················· | ···················· | ···················· | ···················· | ···················· | ···················· |
| V | 6163 | hCoV-19/Japan/PG-8119/2020\|EPI_ISL_687218\|2020-08 | ···················· | ···················· | ···················· | ···················· | ···················· | ···················· |
| V | 6164 | hCoV-19/Japan/PG-8120/2020\|EPI_ISL_687219\|2020-08 | ···················· | ···················· | ···················· | ···················· | ···················· | ···················· |
| V | 6165 | hCoV-19/Japan/PG-8121/2020\|EPI_ISL_687220\|2020-08 | ···················· | ···················· | ···················· | ···················· | ···················· | ···················· |
| V | 6166 | hCoV-19/Japan/PG-8122/2020\|EPI_ISL_687221\|2020-08 | ···················· | ···················· | ···················· | ···················· | ···················· | ···················· |
| V | 6167 | hCoV-19/Japan/PG-8123/2020\|EPI_ISL_687222\|2020-08 | ···················· | ···················· | ···················· | ···················· | ···················· | ···················· |
| V | 6168 | hCoV-19/Japan/PG-8124/2020\|EPI_ISL_687223\|2020-08 | ···················· | ···················· | ···················· | ···················· | ···················· | ···················· |
| V | 6169 | hCoV-19/Japan/PG-8125/2020\|EPI_ISL_687224\|2020-08 | ···················· | ···················· | ···················· | ···················· | ···················· | ···················· |
| V | 6170 | hCoV-19/Japan/PG-8126/2020\|EPI_ISL_687225\|2020-08 | ···················· | ···················· | ···················· | ···················· | ···················· | ···················· |
| V | 6171 | hCoV-19/Japan/PG-8127/2020\|EPI_ISL_687226\|2020-08 | ···················· | ···················· | ···················· | ···················· | ···················· | ···················· |
| V | 6172 | hCoV-19/Japan/PG-8128/2020\|EPI_ISL_687227\|2020-08 | ···················· | ···················· | ···················· | ···················· | ···················· | ···················· |
| V | 6173 | hCoV-19/Japan/PG-8129/2020\|EPI_ISL_687228\|2020-08 | ···················· | ···················· | ···················· | ···················· | ···················· | ···················· |
| V | 6174 | hCoV-19/Japan/PG-8130/2020\|EPI_ISL_687229\|2020-08 | ···················· | ···················· | ···················· | ···················· | ···················· | ···················· |
| V | 6175 | hCoV-19/Japan/PG-8131/2020\|EPI_ISL_687230\|2020-08 | ···················· | ···················· | ···················· | ···················· | ···················· | ···················· |
| V | 6176 | hCoV-19/Japan/PG-8132/2020\|EPI_ISL_687231\|2020-08 | ···················· | ···················· | ···················· | ···················· | ···················· | ···················· |
| V | 6177 | hCoV-19/Japan/PG-8133/2020\|EPI_ISL_687232\|2020-08 | ···················· | ···················· | ···················· | ···················· | ···················· | ···················· |
| V | 6178 | hCoV-19/Japan/PG-8134/2020\|EPI_ISL_687233\|2020-08 | ···················· | ···················· | ···················· | ···················· | ···················· | ···················· |
| V | 6179 | hCoV-19/Japan/PG-8135/2020\|EPI_ISL_687234\|2020-08 | ···················· | ···················· | ···················· | ···················· | ···················· | ···················· |
| V | 6180 | hCoV-19/Japan/PG-8136/2020\|EPI_ISL_687235\|2020-08 | ···················· | ···················· | ···················· | ···················· | ···················· | ···················· |
| V | 6181 | hCoV-19/Japan/PG-8137/2020\|EPI_ISL_687236\|2020-08 | ···················· | ···················· | ···················· | ···················· | ···················· | ···················· |
| V | 6182 | hCoV-19/Japan/PG-8138/2020\|EPI_ISL_687237\|2020-08 | ···················· | ···················· | ···················· | ···················· | ···················· | ···················· |
| V | 6183 | hCoV-19/Japan/PG-8139/2020\|EPI_ISL_687238\|2020-08 | ···················· | ···················· | ···················· | ···················· | ···················· | ···················· |
| V | 6184 | hCoV-19/Japan/PG-8140/2020\|EPI_ISL_687239\|2020-08 | ···················· | ···················· | ···················· | ···················· | ···················· | ···················· |
| V | 6185 | hCoV-19/Japan/PG-8141/2020\|EPI_ISL_687240\|2020-08 | ···················· | ···················· | ···················· | ···················· | ···················· | ···················· |
| V | 6186 | hCoV-19/Japan/PG-8142/2020\|EPI_ISL_687241\|2020-08 | ···················· | ···················· | ···················· | ···················· | ···················· | ···················· |
| V | 6187 | hCoV-19/Japan/PG-8143/2020\|EPI_ISL_687242\|2020-08 | ···················· | ···················· | ···················· | ···················· | ···················· | ···················· |
| V | 6188 | hCoV-19/Japan/PG-8144/2020\|EPI_ISL_687243\|2020-08 | ···················· | ···················· | ···················· | ···················· | ···················· | ···················· |
| V | 6189 | hCoV-19/Japan/PG-8145/2020\|EPI_ISL_687244\|2020-08 | ···················· | ···················· | ···················· | ···················· | ···················· | ···················· |
| V | 6190 | hCoV-19/Japan/PG-8146/2020\|EPI_ISL_687245\|2020-08 | ···················· | ···················· | ···················· | ···················· | ···················· | ···················· |
| V | 6191 | hCoV-19/Japan/PG-8147/2020\|EPI_ISL_687246\|2020-09 | ···················· | ···················· | ···················· | ···················· | ···················· | ···················· |
| V | 6192 | hCoV-19/Japan/PG-8148/2020\|EPI_ISL_687247\|2020-09 | ···················· | ···················· | ···················· | ···················· | ···················· | ···················· |
| V | 6193 | hCoV-19/Japan/PG-8149/2020\|EPI_ISL_687248\|2020-09 | ···················· | ···················· | ···················· | ···················· | ···················· | ···················· |
| V | 6194 | hCoV-19/Japan/PG-8150/2020\|EPI_ISL_687249\|2020-09 | ···················· | ···················· | ···················· | ···················· | ···················· | ···················· |
| V | 6195 | hCoV-19/Japan/PG-8151/2020\|EPI_ISL_687250\|2020-09 | ···················· | ···················· | ···················· | ···················· | ···················· | ···················· |
| V | 6196 | hCoV-19/Japan/PG-8152/2020\|EPI_ISL_687251\|2020-09 | ···················· | ···················· | ···················· | ···················· | ···················· | ···················· |
| V | 6197 | hCoV-19/Japan/PG-8153/2020\|EPI_ISL_687252\|2020-09 | ···················· | ···················· | ···················· | ···················· | ···················· | ···················· |
| V | 6198 | hCoV-19/Japan/PG-8154/2020\|EPI_ISL_687253\|2020-09 | ···················· | ···················· | ···················· | ···················· | ···················· | ···················· |
| V | 6199 | hCoV-19/Japan/PG-8155/2020\|EPI_ISL_687254\|2020-09 | ···················· | ···················· | ···················· | ···················· | ···················· | ···················· |
| V | 6200 | hCoV-19/Japan/PG-8156/2020\|EPI_ISL_687255\|2020-07 | ···················· | ···················· | ···················· | ···················· | ···················· | ···················· |
| V | 6201 | hCoV-19/Japan/PG-8157/2020\|EPI_ISL_687256\|2020-07 | ···················· | ···················· | ···················· | ···················· | ···················· | ···················· |
| V | 6202 | hCoV-19/Japan/PG-8158/2020\|EPI_ISL_687257\|2020-07 | ···················· | ···················· | ···················· | ···················· | ···················· | ···················· |
| V | 6203 | hCoV-19/Japan/PG-8159/2020\|EPI_ISL_687258\|2020-07 | ···················· | ···················· | ···················· | ···················· | ···················· | ···················· |
| V | 6204 | hCoV-19/Japan/PG-8160/2020\|EPI_ISL_687259\|2020-07 | ···················· | ···················· | ···················· | ···················· | ···················· | ···················· |
| V | 6205 | hCoV-19/Japan/PG-8161/2020\|EPI_ISL_687260\|2020-07 | ···················· | ···················· | ···················· | ···················· | ···················· | ···················· |
| V | 6206 | hCoV-19/Japan/PG-8162/2020\|EPI_ISL_687261\|2020-07 | ···················· | ···················· | ···················· | ···················· | ···················· | ···················· |
| V | 6207 | hCoV-19/Japan/PG-8163/2020\|EPI_ISL_687262\|2020-07 | ···················· | ···················· | ···················· | ···················· | ···················· | ···················· |
| V | 6208 | hCoV-19/Japan/PG-8164/2020\|EPI_ISL_687263\|2020-07 | ···················· | ···················· | ···················· | ···················· | ···················· | ···················· |
| V | 6209 | hCoV-19/Japan/PG-8165/2020\|EPI_ISL_687264\|2020-07 | ···················· | ···················· | ···················· | ···················· | ···················· | ···················· |
| V | 6210 | hCoV-19/Japan/PG-8166/2020\|EPI_ISL_687265\|2020-07 | ···················· | ···················· | ···················· | ···················· | ···················· | ···················· |
| V | 6211 | hCoV-19/Japan/PG-8167/2020\|EPI_ISL_687266\|2020-07 | ···················· | ···················· | ···················· | ···················· | ···················· | ···················· |

| | | | | | | | | |
|---|---|---|---|---|---|---|---|---|
| V | 6212 | hCoV-19/Japan/PG-8168/2020\|EPI_ISL_687267\|2020-07 | .................... | .................... | ..................... | .................... | .................... | .................... |
| V | 6213 | hCoV-19/Japan/PG-8169/2020\|EPI_ISL_687268\|2020-07 | .................... | .................... | ..................... | .................... | .................... | .................... |
| V | 6214 | hCoV-19/Japan/PG-8170/2020\|EPI_ISL_687269\|2020-07 | .................... | .................... | ..................... | .................... | .................... | .................... |
| V | 6215 | hCoV-19/Japan/PG-7412/2020\|EPI_ISL_688514\|2020-07 | .................... | .................... | ..................... | .................... | .................... | .................... |
| V | 6216 | hCoV-19/Japan/PG-7413/2020\|EPI_ISL_688515\|2020-07 | .................... | .................... | ..................... | .................... | .................... | .................... |
| V | 6217 | hCoV-19/Japan/PG-7414/2020\|EPI_ISL_688516\|2020-07 | .................... | .................... | ..................... | .................... | .................... | .................... |
| V | 6218 | hCoV-19/Japan/PG-7415/2020\|EPI_ISL_688517\|2020-07 | .................... | .................... | ..................... | .................... | .................... | .................... |
| V | 6219 | hCoV-19/Japan/PG-7416/2020\|EPI_ISL_688518\|2020-07 | .................... | .................... | ..................... | .................... | .................... | .................... |
| V | 6220 | hCoV-19/Japan/PG-7417/2020\|EPI_ISL_688519\|2020-07 | .................... | .................... | ..................... | .................... | .................... | .................... |
| V | 6221 | hCoV-19/Japan/PG-7418/2020\|EPI_ISL_688520\|2020-07 | .................... | .................... | ..................... | .................... | .................... | .................... |
| V | 6222 | hCoV-19/Japan/PG-7419/2020\|EPI_ISL_688521\|2020-07 | .................... | .................... | ..................... | .................... | .................... | .................... |
| V | 6223 | hCoV-19/Japan/PG-7420/2020\|EPI_ISL_688522\|2020-07 | .................... | .................... | ..................... | .................... | .................... | .................... |
| V | 6224 | hCoV-19/Japan/PG-7421/2020\|EPI_ISL_688523\|2020-07 | .................... | .................... | ..................... | .................... | .................... | .................... |
| V | 6225 | hCoV-19/Japan/PG-7422/2020\|EPI_ISL_688524\|2020-07 | .................... | .................... | ..................... | .................... | .................... | .................... |
| V | 6226 | hCoV-19/Japan/PG-7423/2020\|EPI_ISL_688525\|2020-07 | .................... | .................... | ..................... | .................... | .................... | .................... |
| V | 6227 | hCoV-19/Japan/PG-7424/2020\|EPI_ISL_688526\|2020-07 | .................... | .................... | ..................... | .................... | .................... | .................... |
| V | 6228 | hCoV-19/Japan/PG-7425/2020\|EPI_ISL_688527\|2020-07 | .................... | .................... | ..................... | .................... | .................... | .................... |
| V | 6229 | hCoV-19/Japan/PG-7426/2020\|EPI_ISL_688528\|2020-07 | .................... | .................... | ..................... | .................... | .................... | .................... |
| V | 6230 | hCoV-19/Japan/PG-7427/2020\|EPI_ISL_688529\|2020-07 | .................... | .................... | ..................... | .................... | .................... | .................... |
| V | 6231 | hCoV-19/Japan/PG-7428/2020\|EPI_ISL_688530\|2020-07 | .................... | .................... | ..................... | .................... | .................... | .................... |
| V | 6232 | hCoV-19/Japan/PG-7429/2020\|EPI_ISL_688531\|2020-07 | .................... | .................... | ..................... | .................... | .................... | .................... |
| V | 6233 | hCoV-19/Japan/PG-7430/2020\|EPI_ISL_688532\|2020-07 | .................... | .................... | ..................... | .................... | .................... | .................... |
| V | 6234 | hCoV-19/Japan/PG-7431/2020\|EPI_ISL_688533\|2020-07 | .................... | .................... | ..................... | .................... | .................... | .................... |
| V | 6235 | hCoV-19/Japan/PG-7432/2020\|EPI_ISL_688534\|2020-07 | .................... | .................... | ..................... | .................... | .................... | .................... |
| V | 6236 | hCoV-19/Japan/PG-7433/2020\|EPI_ISL_688535\|2020-07 | .................... | .................... | ..................... | .................... | .................... | .................... |
| V | 6237 | hCoV-19/Japan/PG-7434/2020\|EPI_ISL_688536\|2020-07 | .................... | .................... | ..................... | .................... | .................... | .................... |
| V | 6238 | hCoV-19/Japan/PG-7435/2020\|EPI_ISL_688537\|2020-07 | .................... | .................... | ..................... | .................... | .................... | .................... |
| V | 6239 | hCoV-19/Japan/PG-7436/2020\|EPI_ISL_688538\|2020-07 | .................... | .................... | ..................... | .................... | .................... | .................... |
| V | 6240 | hCoV-19/Japan/PG-7437/2020\|EPI_ISL_688539\|2020-07 | .................... | .................... | ..................... | .................... | .................... | .................... |
| V | 6241 | hCoV-19/Japan/PG-7438/2020\|EPI_ISL_688540\|2020-07 | .................... | .................... | ..................... | .................... | .................... | .................... |
| V | 6242 | hCoV-19/Japan/PG-7439/2020\|EPI_ISL_688541\|2020-07 | .................... | .................... | ..................... | .................... | .................... | .................... |
| V | 6243 | hCoV-19/Japan/PG-7440/2020\|EPI_ISL_688542\|2020-07 | .................... | .................... | ..................... | .................... | .................... | .................... |
| V | 6244 | hCoV-19/Japan/PG-7441/2020\|EPI_ISL_688543\|2020-07 | .................... | .................... | ..................... | .................... | .................... | .................... |
| V | 6245 | hCoV-19/Japan/PG-7442/2020\|EPI_ISL_688544\|2020-07 | .................... | .................... | ..................... | .................... | .................... | .................... |
| V | 6246 | hCoV-19/Japan/PG-7443/2020\|EPI_ISL_688545\|2020-07 | .................... | .................... | ..................... | .................... | .................... | .................... |
| V | 6247 | hCoV-19/Japan/PG-7444/2020\|EPI_ISL_688546\|2020-07 | .................... | .................... | ..................... | .................... | .................... | .................... |
| V | 6248 | hCoV-19/Japan/PG-7445/2020\|EPI_ISL_688547\|2020-07 | .................... | .................... | ..................... | .................... | .................... | .................... |
| V | 6249 | hCoV-19/Japan/PG-7446/2020\|EPI_ISL_688548\|2020-07 | .................... | .................... | ..................... | .................... | .................... | .................... |
| V | 6250 | hCoV-19/Japan/PG-7447/2020\|EPI_ISL_688549\|2020-07 | .................... | .................... | ..................... | .................... | .................... | .................... |
| V | 6251 | hCoV-19/Japan/PG-7448/2020\|EPI_ISL_688550\|2020-07 | .................... | .................... | ..................... | .................... | .................... | .................... |
| V | 6252 | hCoV-19/Japan/PG-7449/2020\|EPI_ISL_688551\|2020-07 | .................... | .................... | ..................... | .................... | .................... | .................... |
| V | 6253 | hCoV-19/Japan/PG-7450/2020\|EPI_ISL_688552\|2020-07 | .................... | .................... | ..................... | .................... | .................... | .................... |
| V | 6254 | hCoV-19/Japan/PG-7451/2020\|EPI_ISL_688553\|2020-07 | .................... | .................... | ..................... | .................... | .................... | .................... |
| V | 6255 | hCoV-19/Japan/PG-7452/2020\|EPI_ISL_688554\|2020-07 | .................... | .................... | ..................... | .................... | .................... | .................... |
| V | 6256 | hCoV-19/Japan/PG-7453/2020\|EPI_ISL_688555\|2020-07 | .................... | .................... | ..................... | .................... | .................... | .................... |
| V | 6257 | hCoV-19/Japan/PG-7454/2020\|EPI_ISL_688556\|2020-07 | .................... | .................... | ..................... | .................... | .................... | .................... |
| V | 6258 | hCoV-19/Japan/PG-7455/2020\|EPI_ISL_688557\|2020-07 | .................... | .................... | ..................... | .................... | .................... | .................... |
| V | 6259 | hCoV-19/Japan/PG-7456/2020\|EPI_ISL_688558\|2020-07 | .................... | .................... | ..................... | .................... | .................... | .................... |
| V | 6260 | hCoV-19/Japan/PG-7457/2020\|EPI_ISL_688559\|2020-07 | .................... | .................... | ..................... | .................... | .................... | .................... |
| V | 6261 | hCoV-19/Japan/PG-7458/2020\|EPI_ISL_688560\|2020-07 | .................... | .................... | ..................... | .................... | .................... | .................... |
| V | 6262 | hCoV-19/Japan/PG-7459/2020\|EPI_ISL_688561\|2020-07 | .................... | .................... | ..................... | .................... | .................... | .................... |
| V | 6263 | hCoV-19/Japan/PG-7460/2020\|EPI_ISL_688562\|2020-07 | .................... | .................... | ..................... | .................... | .................... | .................... |
| V | 6264 | hCoV-19/Japan/PG-7461/2020\|EPI_ISL_688563\|2020-07 | .................... | .................... | ..................... | .................... | .................... | .................... |
| V | 6265 | hCoV-19/Japan/PG-7462/2020\|EPI_ISL_688564\|2020-07 | .................... | .................... | ..................... | .................... | .................... | .................... |
| V | 6266 | hCoV-19/Japan/PG-7463/2020\|EPI_ISL_688565\|2020-07 | .................... | .................... | ..................... | .................... | .................... | .................... |
| V | 6267 | hCoV-19/Japan/PG-7464/2020\|EPI_ISL_688566\|2020-07 | .................... | .................... | ..................... | .................... | .................... | .................... |
| V | 6268 | hCoV-19/Japan/PG-7465/2020\|EPI_ISL_688567\|2020-08 | .................... | .................... | ..................... | .................... | .................... | .................... |
| V | 6269 | hCoV-19/Japan/PG-7466/2020\|EPI_ISL_688568\|2020-07 | .................... | .................... | ..................... | .................... | .................... | .................... |
| V | 6270 | hCoV-19/Japan/PG-7467/2020\|EPI_ISL_688569\|2020-07 | .................... | .................... | ..................... | .................... | .................... | .................... |
| V | 6271 | hCoV-19/Japan/PG-7468/2020\|EPI_ISL_688570\|2020-07 | .................... | .................... | ..................... | .................... | .................... | .................... |
| V | 6272 | hCoV-19/Japan/PG-7469/2020\|EPI_ISL_688571\|2020-07 | .................... | .................... | ..................... | .................... | .................... | .................... |
| V | 6273 | hCoV-19/Japan/PG-7470/2020\|EPI_ISL_688572\|2020-07 | .................... | .................... | ..................... | .................... | .................... | .................... |
| V | 6274 | hCoV-19/Japan/PG-7471/2020\|EPI_ISL_688573\|2020-07 | .................... | .................... | ..................... | .................... | .................... | .................... |
| V | 6275 | hCoV-19/Japan/PG-7472/2020\|EPI_ISL_688574\|2020-07 | .................... | .................... | ..................... | .................... | .................... | .................... |
| V | 6276 | hCoV-19/Japan/PG-7473/2020\|EPI_ISL_688575\|2020-07 | .................... | .................... | ..................... | .................... | .................... | .................... |
| V | 6277 | hCoV-19/Japan/PG-7474/2020\|EPI_ISL_688576\|2020-07 | .................... | .................... | ..................... | .................... | .................... | .................... |
| V | 6278 | hCoV-19/Japan/PG-7475/2020\|EPI_ISL_688577\|2020-07 | .................... | .................... | ..................... | .................... | .................... | .................... |
| V | 6279 | hCoV-19/Japan/PG-7476/2020\|EPI_ISL_688578\|2020-07 | .................... | .................... | ..................... | .................... | .................... | .................... |
| V | 6280 | hCoV-19/Japan/PG-7477/2020\|EPI_ISL_688579\|2020-07 | .................... | .................... | ..................... | .................... | .................... | .................... |
| V | 6281 | hCoV-19/Japan/PG-7478/2020\|EPI_ISL_688580\|2020-07 | .................... | .................... | ..................... | .................... | .................... | .................... |
| V | 6282 | hCoV-19/Japan/PG-7479/2020\|EPI_ISL_688581\|2020-07 | .................... | .................... | ..................... | .................... | .................... | .................... |
| V | 6283 | hCoV-19/Japan/PG-7480/2020\|EPI_ISL_688582\|2020-07 | .................... | .................... | ..................... | .................... | .................... | .................... |
| V | 6284 | hCoV-19/Japan/PG-7481/2020\|EPI_ISL_688583\|2020-08 | .................... | .................... | ..................... | .................... | .................... | .................... |
| V | 6285 | hCoV-19/Japan/PG-7482/2020\|EPI_ISL_688584\|2020-08 | .................... | .................... | ..................... | .................... | .................... | .................... |
| V | 6286 | hCoV-19/Japan/PG-7483/2020\|EPI_ISL_688585\|2020-08 | .................... | .................... | ..................... | .................... | .................... | .................... |
| V | 6287 | hCoV-19/Japan/PG-7484/2020\|EPI_ISL_688586\|2020-08 | .................... | .................... | ..................... | .................... | .................... | .................... |
| V | 6288 | hCoV-19/Japan/PG-7485/2020\|EPI_ISL_688587\|2020-08 | .................... | .................... | ..................... | .................... | .................... | .................... |
| V | 6289 | hCoV-19/Japan/PG-7486/2020\|EPI_ISL_688588\|2020-08 | .................... | .................... | ..................... | .................... | .................... | .................... |
| V | 6290 | hCoV-19/Japan/PG-7487/2020\|EPI_ISL_688589\|2020-08 | .................... | .................... | ..................... | .................... | .................... | .................... |
| V | 6291 | hCoV-19/Japan/PG-7488/2020\|EPI_ISL_688590\|2020-08 | .................... | .................... | ..................... | .................... | .................... | .................... |

| | | | | | | | |
|---|---|---|---|---|---|---|---|
| V | 6292 hCoV-19/Japan/PG-7489/2020\|EPI_ISL_688591\|2020-08 | ···················· | ···················· | ····················· | ···················· | ···················· | ···················· |
| V | 6293 hCoV-19/Japan/PG-7490/2020\|EPI_ISL_688592\|2020-08 | ···················· | ···················· | ····················· | ···················· | ···················· | ···················· |
| V | 6294 hCoV-19/Japan/PG-7491/2020\|EPI_ISL_688593\|2020-08 | ···················· | ···················· | ····················· | ···················· | ···················· | ···················· |
| V | 6295 hCoV-19/Japan/PG-7492/2020\|EPI_ISL_688594\|2020-08 | ···················· | ···················· | ····················· | ···················· | ···················· | ···················· |
| V | 6296 hCoV-19/Japan/PG-7493/2020\|EPI_ISL_688595\|2020-08 | ···················· | ···················· | ····················· | ···················· | ···················· | ···················· |
| V | 6297 hCoV-19/Japan/PG-7494/2020\|EPI_ISL_688596\|2020-08 | ···················· | ···················· | ····················· | ···················· | ···················· | ···················· |
| V | 6298 hCoV-19/Japan/PG-7495/2020\|EPI_ISL_688597\|2020-08 | ···················· | ···················· | ····················· | ···················· | ···················· | ···················· |
| V | 6299 hCoV-19/Japan/PG-7496/2020\|EPI_ISL_688598\|2020-08 | ···················· | ···················· | ····················· | ···················· | ···················· | ···················· |
| V | 6300 hCoV-19/Japan/PG-7497/2020\|EPI_ISL_688599\|2020-08 | ···················· | ···················· | ····················· | ···················· | ···················· | ···················· |
| V | 6301 hCoV-19/Japan/PG-7498/2020\|EPI_ISL_688600\|2020-08 | ···················· | ···················· | ····················· | ···················· | ···················· | ···················· |
| V | 6302 hCoV-19/Japan/PG-7499/2020\|EPI_ISL_688601\|2020-08 | ···················· | ···················· | ····················· | ···················· | ···················· | ···················· |
| V | 6303 hCoV-19/Japan/PG-7500/2020\|EPI_ISL_688602\|2020-08 | ···················· | ···················· | ····················· | ···················· | ···················· | ···················· |
| V | 6304 hCoV-19/Japan/PG-7501/2020\|EPI_ISL_688603\|2020-08 | ···················· | ···················· | ····················· | ···················· | ···················· | ···················· |
| V | 6305 hCoV-19/Japan/PG-7502/2020\|EPI_ISL_688604\|2020-08 | ···················· | ···················· | ····················· | ···················· | ···················· | ···················· |
| V | 6306 hCoV-19/Japan/PG-7503/2020\|EPI_ISL_688605\|2020-07 | ···················· | ···················· | ····················· | ···················· | ···················· | ···················· |
| V | 6307 hCoV-19/Japan/PG-7504/2020\|EPI_ISL_688606\|2020-07 | ···················· | ···················· | ····················· | ···················· | ···················· | ···················· |
| V | 6308 hCoV-19/Japan/PG-7505/2020\|EPI_ISL_688607\|2020-07 | ···················· | ···················· | ····················· | ···················· | ···················· | ···················· |
| V | 6309 hCoV-19/Japan/PG-7506/2020\|EPI_ISL_688608\|2020-07 | ···················· | ···················· | ····················· | ···················· | ···················· | ···················· |
| V | 6310 hCoV-19/Japan/PG-7507/2020\|EPI_ISL_688609\|2020-07 | ···················· | ···················· | ····················· | ···················· | ···················· | ···················· |
| V | 6311 hCoV-19/Japan/PG-7508/2020\|EPI_ISL_688610\|2020-07 | ···················· | ···················· | ····················· | ···················· | ···················· | ···················· |
| V | 6312 hCoV-19/Japan/PG-7509/2020\|EPI_ISL_688611\|2020-07 | ···················· | ···················· | ····················· | ···················· | ···················· | ···················· |
| V | 6313 hCoV-19/Japan/PG-5801/2020\|EPI_ISL_688612\|2020-08 | ···················· | ···················· | ····················· | ···················· | ···················· | ···················· |
| V | 6314 hCoV-19/Japan/PG-5865/2020\|EPI_ISL_688613\|2020-08 | ···················· | ···················· | ····················· | ···················· | ···················· | ···················· |
| V | 6315 hCoV-19/Japan/PG-6445/2020\|EPI_ISL_688614\|2020-09 | ···················· | ···················· | ····················· | ···················· | ···················· | ···················· |
| V | 6316 hCoV-19/Japan/PG-5744/2020\|EPI_ISL_688616\|2020-08 | ···················· | ···················· | ····················· | ···················· | ···················· | ···················· |
| V | 6317 hCoV-19/Japan/PG-5984/2020\|EPI_ISL_688617\|2020-08 | ···················· | ···················· | ····················· | ···················· | ···················· | ···················· |
| V | 6318 hCoV-19/Japan/PG-5985/2020\|EPI_ISL_688618\|2020-08 | ···················· | ···················· | ····················· | ···················· | ···················· | ···················· |
| V | 6319 hCoV-19/Japan/PG-6378/2020\|EPI_ISL_688619\|2020-08 | ···················· | ···················· | ····················· | ···················· | ···················· | ···················· |
| V | 6320 hCoV-19/Japan/PG-6383/2020\|EPI_ISL_688620\|2020-08 | ···················· | ···················· | ····················· | ···················· | ···················· | ···················· |
| V | 6321 hCoV-19/Japan/PG-6388/2020\|EPI_ISL_688621\|2020-08 | ···················· | ···················· | ····················· | ···················· | ···················· | ···················· |
| V | 6322 hCoV-19/Japan/PG-6389/2020\|EPI_ISL_688622\|2020-08 | ···················· | ···················· | ····················· | ···················· | ···················· | ···················· |
| V | 6323 hCoV-19/Japan/PG-6390/2020\|EPI_ISL_688623\|2020-08 | ···················· | ···················· | ····················· | ···················· | ···················· | ···················· |
| V | 6324 hCoV-19/Japan/PG-4708/2020\|EPI_ISL_689845\|2020-08 | ···················· | ···················· | ····················· | ···················· | ···················· | ···················· |
| V | 6325 hCoV-19/Japan/PG-4709/2020\|EPI_ISL_689846\|2020-07 | ···················· | ···················· | ····················· | ···················· | ···················· | ···················· |
| V | 6326 hCoV-19/Japan/PG-4710/2020\|EPI_ISL_689847\|2020-08 | ···················· | ···················· | ····················· | ···················· | ···················· | ···················· |
| V | 6327 hCoV-19/Japan/PG-4711/2020\|EPI_ISL_689848\|2020-08 | ···················· | ···················· | ····················· | ···················· | ···················· | ···················· |
| V | 6328 hCoV-19/Japan/PG-4712/2020\|EPI_ISL_689849\|2020-08 | ···················· | ···················· | ····················· | ···················· | ···················· | ···················· |
| V | 6329 hCoV-19/Japan/PG-4713/2020\|EPI_ISL_689850\|2020-08 | ···················· | ···················· | ····················· | ···················· | ···················· | ···················· |
| V | 6330 hCoV-19/Japan/PG-4715/2020\|EPI_ISL_689851\|2020-07 | ···················· | ···················· | ····················· | ···················· | ···················· | ···················· |
| V | 6331 hCoV-19/Japan/PG-4716/2020\|EPI_ISL_689852\|2020-07 | ···················· | ···················· | ····················· | ···················· | ···················· | ···················· |
| V | 6332 hCoV-19/Japan/PG-4717/2020\|EPI_ISL_689853\|2020-07 | ···················· | ···················· | ····················· | ···················· | ···················· | ···················· |
| V | 6333 hCoV-19/Japan/PG-4718/2020\|EPI_ISL_689854\|2020-07 | ···················· | ···················· | ····················· | ···················· | ···················· | ···················· |
| V | 6334 hCoV-19/Japan/PG-4719/2020\|EPI_ISL_689855\|2020-07 | ···················· | ···················· | ····················· | ···················· | ···················· | ···················· |
| V | 6335 hCoV-19/Japan/PG-4720/2020\|EPI_ISL_689856\|2020-07 | ···················· | ···················· | ····················· | ···················· | ···················· | ···················· |
| V | 6336 hCoV-19/Japan/PG-4721/2020\|EPI_ISL_689857\|2020-07 | ···················· | ···················· | ····················· | ···················· | ···················· | ···················· |
| V | 6337 hCoV-19/Japan/PG-4722/2020\|EPI_ISL_689858\|2020-07 | ···················· | ···················· | ····················· | ···················· | ···················· | ···················· |
| V | 6338 hCoV-19/Japan/PG-4723/2020\|EPI_ISL_689859\|2020-07 | ···················· | ···················· | ····················· | ···················· | ···················· | ···················· |
| V | 6339 hCoV-19/Japan/PG-4724/2020\|EPI_ISL_689860\|2020-07 | ···················· | ···················· | ····················· | ···················· | ···················· | ···················· |
| V | 6340 hCoV-19/Japan/PG-4725/2020\|EPI_ISL_689861\|2020-07 | ···················· | ···················· | ····················· | ···················· | ···················· | ···················· |
| V | 6341 hCoV-19/Japan/PG-4726/2020\|EPI_ISL_689862\|2020-07 | ···················· | ···················· | ····················· | ···················· | ···················· | ···················· |
| V | 6342 hCoV-19/Japan/PG-4727/2020\|EPI_ISL_689863\|2020-07 | ···················· | ···················· | ····················· | ···················· | ···················· | ···················· |
| V | 6343 hCoV-19/Japan/PG-4728/2020\|EPI_ISL_689864\|2020-07 | ···················· | ···················· | ····················· | ···················· | ···················· | ···················· |
| V | 6344 hCoV-19/Japan/PG-4729/2020\|EPI_ISL_689865\|2020-07 | ···················· | ···················· | ····················· | ···················· | ···················· | ···················· |
| V | 6345 hCoV-19/Japan/PG-4730/2020\|EPI_ISL_689866\|2020-07 | ···················· | ···················· | ····················· | ···················· | ···················· | ···················· |
| V | 6346 hCoV-19/Japan/PG-4731/2020\|EPI_ISL_689867\|2020-07 | ···················· | ···················· | ····················· | ···················· | ···················· | ···················· |
| V | 6347 hCoV-19/Japan/PG-4732/2020\|EPI_ISL_689868\|2020-07 | ···················· | ···················· | ····················· | ···················· | ···················· | ···················· |
| V | 6348 hCoV-19/Japan/PG-4733/2020\|EPI_ISL_689869\|2020-07 | ···················· | ···················· | ····················· | ···················· | ···················· | ···················· |
| V | 6349 hCoV-19/Japan/PG-2698/2020\|EPI_ISL_690830\|2020-04 | ···················· | ···················· | ····················· | ···················· | ···················· | ···················· |
| V | 6350 hCoV-19/Japan/PG-2699/2020\|EPI_ISL_690831\|2020-04 | ···················· | ·A·················· | ····················· | ···················· | ···················· | ···················· |
| V | 6351 hCoV-19/Japan/PG-2700/2020\|EPI_ISL_690832\|2020-04 | ···················· | ···················· | ····················· | ···················· | ···················· | ···················· |
| V | 6352 hCoV-19/Japan/PG-2701/2020\|EPI_ISL_690833\|2020-04 | ···················· | ···················· | ····················· | ···················· | ···················· | ···················· |
| V | 6353 hCoV-19/Japan/PG-2702/2020\|EPI_ISL_690834\|2020-04 | ···················· | ···················· | ····················· | ···················· | ···················· | ···················· |
| V | 6354 hCoV-19/Japan/PG-2703/2020\|EPI_ISL_690835\|2020-05 | ···················· | ···················· | ····················· | ···················· | ···················· | ···················· |
| V | 6355 hCoV-19/Japan/PG-2704/2020\|EPI_ISL_690836\|2020-05 | ···················· | ···················· | ····················· | ···················· | ···················· | ···················· |
| V | 6356 hCoV-19/Japan/PG-2705/2020\|EPI_ISL_690837\|2020-04 | ···················· | ···················· | ····················· | ···················· | ···················· | ···················· |
| V | 6357 hCoV-19/Japan/PG-2706/2020\|EPI_ISL_690838\|2020-04 | ···················· | ···················· | ····················· | ···················· | ···················· | ···················· |
| V | 6358 hCoV-19/Japan/PG-2707/2020\|EPI_ISL_690839\|2020-04 | ···················· | ···················· | ····················· | ···················· | ···················· | ···················· |
| V | 6359 hCoV-19/Japan/PG-2708/2020\|EPI_ISL_690840\|2020-04 | ···················· | ···················· | ····················· | ···················· | ···················· | ···················· |
| V | 6360 hCoV-19/Japan/PG-2709/2020\|EPI_ISL_690841\|2020-04 | ···················· | ···················· | ····················· | ···················· | ···················· | ···················· |
| V | 6361 hCoV-19/Japan/PG-2710/2020\|EPI_ISL_690842\|2020-05 | ···················· | ···················· | ····················· | ···················· | ···················· | ···················· |
| V | 6362 hCoV-19/Japan/PG-2711/2020\|EPI_ISL_690843\|2020-05 | ···················· | ···················· | ····················· | ···················· | ···················· | ···················· |
| V | 6363 hCoV-19/Japan/PG-2712/2020\|EPI_ISL_690844\|2020-06 | ···················· | ···················· | ····················· | ···················· | ···················· | ···················· |
| V | 6364 hCoV-19/Japan/PG-2713/2020\|EPI_ISL_690845\|2020-03 | ···················· | ···················· | ····················· | ···················· | ···················· | ···················· |
| V | 6365 hCoV-19/Japan/PG-2714/2020\|EPI_ISL_690846\|2020-03 | ···················· | ···················· | ····················· | ···················· | ···················· | ···················· |
| V | 6366 hCoV-19/Japan/PG-2715/2020\|EPI_ISL_690847\|2020-03 | ···················· | ···················· | ····················· | ···················· | ···················· | ···················· |
| V | 6367 hCoV-19/Japan/PG-2716/2020\|EPI_ISL_690848\|2020-03 | ···················· | ···················· | ····················· | ···················· | ···················· | ···················· |
| V | 6368 hCoV-19/Japan/PG-2717/2020\|EPI_ISL_690849\|2020-03 | ···················· | ···················· | ····················· | ···················· | ···················· | ···················· |
| V | 6369 hCoV-19/Japan/PG-2718/2020\|EPI_ISL_690850\|2020-03 | ···················· | ···················· | ····················· | ···················· | ···················· | ···················· |
| V | 6370 hCoV-19/Japan/PG-2720/2020\|EPI_ISL_690852\|2020-03 | ···················· | ···················· | ····················· | ···················· | ···················· | ···················· |
| V | 6371 hCoV-19/Japan/PG-2721/2020\|EPI_ISL_690853\|2020-03 | ···················· | ···················· | ····················· | ···················· | ···················· | ···················· |

| | | | | | | | |
|---|---|---|---|---|---|---|---|
| V | 6372 hCoV-19/Japan/PG-2722/2020\|EPI_ISL_690854\|2020-03 | ···················· | ···················· | ····················· | ···················· | ···················· | ···················· |
| V | 6373 hCoV-19/Japan/PG-2723/2020\|EPI_ISL_690855\|2020-03 | ···················· | ···················· | ····················· | ···················· | ···················· | ···················· |
| V | 6374 hCoV-19/Japan/PG-2724/2020\|EPI_ISL_690856\|2020-03 | ···················· | ···················· | ····················· | ···················· | ···················· | ···················· |
| V | 6375 hCoV-19/Japan/PG-2725/2020\|EPI_ISL_690857\|2020-03 | ···················· | ···················· | ····················· | ···················· | ···················· | ···················· |
| V | 6376 hCoV-19/Japan/PG-2726/2020\|EPI_ISL_690858\|2020-03 | ···················· | ···················· | ····················· | ···················· | ···················· | ···················· |
| V | 6377 hCoV-19/Japan/PG-2727/2020\|EPI_ISL_690859\|2020-03 | ···················· | ···················· | ····················· | ···················· | ···················· | ···················· |
| V | 6378 hCoV-19/USA/MA-MASPHL-00460/2020\|EPI_ISL_693102\|20 | ···················· | ···················· | ····················· | ···················· | ···················· | ···················· |
| V | 6379 hCoV-19/USA/MA-MASPHL-00459/2020\|EPI_ISL_693103\|20 | ···················· | ···················· | ····················· | ···················· | ···················· | ···················· |
| V | 6380 hCoV-19/USA/MA-MASPHL-00457/2020\|EPI_ISL_693105\|20 | ···················· | ···················· | ····················· | ···················· | ···················· | ···················· |
| V | 6381 hCoV-19/USA/MA-MASPHL-00456/2020\|EPI_ISL_693106\|20 | ···················· | ···················· | ····················· | ···················· | ···················· | ···················· |
| V | 6382 hCoV-19/USA/MA-MASPHL-00454/2020\|EPI_ISL_693107\|20 | ···················· | ···················· | ····················· | ···················· | ···················· | ···················· |
| V | 6383 hCoV-19/USA/MA-MASPHL-00451/2020\|EPI_ISL_693109\|20 | ···················· | ···················· | ····················· | ···················· | ···················· | ···················· |
| V | 6384 hCoV-19/USA/MA-MASPHL-00450/2020\|EPI_ISL_693110\|20 | ···················· | ···················· | ····················· | ···················· | ···················· | ···················· |
| V | 6385 hCoV-19/USA/MA-MASPHL-00446/2020\|EPI_ISL_693113\|20 | ···················· | ···················· | ····················· | ···················· | ···················· | ···················· |
| V | 6386 hCoV-19/USA/MA-MASPHL-00445/2020\|EPI_ISL_693114\|20 | ···················· | ···················· | ····················· | ···················· | ···················· | ···················· |
| V | 6387 hCoV-19/USA/MA-MASPHL-00444/2020\|EPI_ISL_693115\|20 | ···················· | ···················· | ····················· | ···················· | ···················· | ···················· |
| V | 6388 hCoV-19/USA/MA-MASPHL-00437/2020\|EPI_ISL_693120\|20 | ···················· | ···················· | ····················· | ···················· | ···················· | ···················· |
| V | 6389 hCoV-19/USA/MA-MASPHL-00436/2020\|EPI_ISL_693121\|20 | ···················· | ···················· | ····················· | ···················· | ···················· | ···················· |
| V | 6390 hCoV-19/USA/MA-MASPHL-00429/2020\|EPI_ISL_693124\|20 | ···················· | ···················· | ····················· | ···················· | ···················· | ···················· |
| V | 6391 hCoV-19/USA/MA-MASPHL-00428/2020\|EPI_ISL_693125\|20 | ···················· | ···················· | ····················· | ···················· | ···················· | ···················· |
| V | 6392 hCoV-19/USA/MA-MASPHL-00427/2020\|EPI_ISL_693126\|20 | ···················· | ···················· | ····················· | ···················· | ···················· | ···················· |
| V | 6393 hCoV-19/USA/MA-MASPHL-00426/2020\|EPI_ISL_693127\|20 | ···················· | ···················· | ····················· | ···················· | ···················· | ···················· |
| V | 6394 hCoV-19/USA/MA-MASPHL-00425/2020\|EPI_ISL_693128\|20 | ···················· | ···················· | ····················· | ···················· | ···················· | ···················· |
| V | 6395 hCoV-19/USA/MA-MASPHL-00421/2020\|EPI_ISL_693130\|20 | ···················· | ···················· | ····················· | ···················· | ···················· | ···················· |
| V | 6396 hCoV-19/USA/MA-MASPHL-00420/2020\|EPI_ISL_693131\|20 | ···················· | ···················· | ····················· | ···················· | ···················· | ···················· |
| V | 6397 hCoV-19/USA/MA-MASPHL-00417/2020\|EPI_ISL_693134\|20 | ···················· | ···················· | ····················· | ···················· | ···················· | ···················· |
| V | 6398 hCoV-19/USA/MA-MASPHL-00412/2020\|EPI_ISL_693137\|20 | ···················· | ···················· | ····················· | ···················· | ···················· | ···················· |
| V | 6399 hCoV-19/USA/MA-MASPHL-00403/2020\|EPI_ISL_693143\|20 | ···················· | ···················· | ····················· | ···················· | ···················· | ···················· |
| V | 6400 hCoV-19/USA/MA-MASPHL-00402/2020\|EPI_ISL_693144\|20 | ···················· | ···················· | ····················· | ···················· | ···················· | ···················· |
| V | 6401 hCoV-19/Japan/PG-1346/2020\|EPI_ISL_684954\|2020-04 | ···················· | ···················· | ····················· | ···················· | ···················· | ···················· |
| V | 6402 hCoV-19/Japan/PG-1347/2020\|EPI_ISL_684955\|2020-04 | ···················· | ···················· | ····················· | ···················· | ···················· | ···················· |
| V | 6403 hCoV-19/Japan/PG-1348/2020\|EPI_ISL_684956\|2020-04 | ···················· | ···················· | ····················· | ···················· | ····C··············· | ···················· |
| V | 6404 hCoV-19/Japan/PG-1349/2020\|EPI_ISL_684957\|2020-04 | ···················· | ···················· | ····················· | ···················· | ···················· | ···················· |
| V | 6405 hCoV-19/Japan/PG-1350/2020\|EPI_ISL_684958\|2020-04 | ···················· | ···················· | ····················· | ···················· | ···················· | ···················· |
| V | 6406 hCoV-19/Japan/PG-1351/2020\|EPI_ISL_684959\|2020-04 | ···················· | ···················· | ····················· | ···················· | ···················· | ···················· |
| V | 6407 hCoV-19/Japan/PG-1352/2020\|EPI_ISL_684960\|2020-04 | ···················· | ···················· | ····················· | ···················· | ···················· | ···················· |
| V | 6408 hCoV-19/Japan/PG-1353/2020\|EPI_ISL_684961\|2020-04 | ···················· | ···················· | ····················· | ···················· | ···················· | ···················· |
| V | 6409 hCoV-19/Japan/PG-1354/2020\|EPI_ISL_684962\|2020-04 | ···················· | ···················· | ····················· | ···················· | ···················· | ···················· |
| V | 6410 hCoV-19/Japan/PG-1355/2020\|EPI_ISL_684963\|2020-04 | ···················· | ···················· | ····················· | ···················· | ···················· | ···················· |
| V | 6411 hCoV-19/Japan/PG-1357/2020\|EPI_ISL_684965\|2020-04 | ···················· | ···················· | ····················· | ···················· | ···················· | ···················· |
| V | 6412 hCoV-19/Japan/PG-1358/2020\|EPI_ISL_684966\|2020-04 | ···················· | ···················· | ····················· | ···················· | ···················· | ···················· |
| V | 6413 hCoV-19/Japan/PG-1359/2020\|EPI_ISL_684967\|2020-04 | ···················· | ···················· | ····················· | ···················· | ···················· | ···················· |
| V | 6414 hCoV-19/Japan/PG-1360/2020\|EPI_ISL_684968\|2020-04 | ···················· | ···················· | ····················· | ···················· | ···················· | ···················· |
| V | 6415 hCoV-19/Japan/PG-1361/2020\|EPI_ISL_684969\|2020-04 | ···················· | ···················· | ····················· | ···················· | ···················· | ···················· |
| V | 6416 hCoV-19/Japan/PG-1362/2020\|EPI_ISL_684970\|2020-04 | ···················· | ···················· | ····················· | ···················· | ···················· | ···················· |
| V | 6417 hCoV-19/Japan/PG-1363/2020\|EPI_ISL_684971\|2020-04 | ···················· | ···················· | ····················· | ···················· | ···················· | ···················· |
| V | 6418 hCoV-19/Japan/PG-1364/2020\|EPI_ISL_684972\|2020-04 | ···················· | ···················· | ····················· | ···················· | ···················· | ···················· |
| V | 6419 hCoV-19/Japan/PG-1365/2020\|EPI_ISL_684973\|2020-04 | ···················· | ···················· | ····················· | ···················· | ···················· | ···················· |
| V | 6420 hCoV-19/Japan/PG-1366/2020\|EPI_ISL_684974\|2020-04 | ···················· | ···················· | ····················· | ···················· | ···················· | ···················· |
| V | 6421 hCoV-19/Japan/PG-1367/2020\|EPI_ISL_684975\|2020-04 | ···················· | ···················· | ····················· | ···················· | ···················· | ···················· |
| V | 6422 hCoV-19/Japan/PG-1368/2020\|EPI_ISL_684976\|2020-04 | ···················· | ···················· | ····················· | ···················· | ···················· | ···················· |
| V | 6423 hCoV-19/Japan/PG-1370/2020\|EPI_ISL_684977\|2020-04 | ···················· | ···················· | ····················· | ···················· | ···················· | ···················· |
| V | 6424 hCoV-19/Japan/PG-1371/2020\|EPI_ISL_684978\|2020-04 | ···················· | ···················· | ····················· | ···················· | ···················· | ···················· |
| V | 6425 hCoV-19/Japan/PG-1372/2020\|EPI_ISL_684979\|2020-04 | ···················· | ···················· | ····················· | ···················· | ···················· | ···················· |
| V | 6426 hCoV-19/Japan/PG-1373/2020\|EPI_ISL_684980\|2020-04 | ···················· | ···················· | ····················· | ···················· | ···················· | ···················· |
| V | 6427 hCoV-19/Japan/PG-1374/2020\|EPI_ISL_684981\|2020-04 | ···················· | ···················· | ····················· | ···················· | ···················· | ···················· |
| V | 6428 hCoV-19/Japan/PG-1375/2020\|EPI_ISL_684982\|2020-04 | ···················· | ···················· | ····················· | ···················· | ···················· | ···················· |
| V | 6429 hCoV-19/Japan/PG-1376/2020\|EPI_ISL_684983\|2020-04 | ···················· | ···················· | ····················· | ···················· | ···················· | ···················· |
| V | 6430 hCoV-19/Japan/PG-1377/2020\|EPI_ISL_684984\|2020-04 | ···················· | ···················· | ····················· | ···················· | ···················· | ···················· |
| V | 6431 hCoV-19/Japan/PG-1378/2020\|EPI_ISL_684985\|2020-04 | ···················· | ···················· | ····················· | ···················· | ···················· | ···················· |
| V | 6432 hCoV-19/Japan/PG-1379/2020\|EPI_ISL_684986\|2020-04 | ···················· | ···················· | ····················· | ···················· | ···················· | ···················· |
| V | 6433 hCoV-19/Japan/PG-1380/2020\|EPI_ISL_684987\|2020-04 | ···················· | ···················· | ····················· | ···················· | ···················· | ···················· |
| V | 6434 hCoV-19/Japan/PG-1381/2020\|EPI_ISL_684988\|2020-04 | ···················· | ···················· | ····················· | ···················· | ···················· | ···················· |
| V | 6435 hCoV-19/Japan/PG-1382/2020\|EPI_ISL_684989\|2020-04 | ···················· | ···················· | ····················· | ···················· | ···················· | ···················· |
| V | 6436 hCoV-19/Japan/PG-1383/2020\|EPI_ISL_684990\|2020-04 | ···················· | ···················· | ····················· | ···················· | ···················· | ···················· |
| V | 6437 hCoV-19/Japan/PG-1384/2020\|EPI_ISL_684991\|2020-04 | ···················· | ···················· | ····················· | ···················· | ···················· | ···················· |
| V | 6438 hCoV-19/Japan/PG-1385/2020\|EPI_ISL_684992\|2020-04 | ···················· | ···················· | ····················· | ···················· | ···················· | ···················· |
| V | 6439 hCoV-19/Japan/PG-1386/2020\|EPI_ISL_684993\|2020-04 | ···················· | ···················· | ····················· | ···················· | ···················· | ···················· |
| V | 6440 hCoV-19/Japan/PG-1387/2020\|EPI_ISL_684994\|2020-04 | ···················· | ···················· | ····················· | ···················· | ···················· | ···················· |
| V | 6441 hCoV-19/Japan/PG-1388/2020\|EPI_ISL_684995\|2020-04 | ···················· | ···················· | ····················· | ···················· | ···················· | ···················· |
| V | 6442 hCoV-19/Japan/PG-1389/2020\|EPI_ISL_684996\|2020-04 | ···················· | ···················· | ····················· | ···················· | ···················· | ···················· |
| V | 6443 hCoV-19/Japan/PG-1390/2020\|EPI_ISL_684997\|2020-04 | ···················· | ···················· | ····················· | ···················· | ···················· | ···················· |
| V | 6444 hCoV-19/Japan/PG-1391/2020\|EPI_ISL_684998\|2020-04 | ···················· | ···················· | ····················· | ···················· | ···················· | ···················· |
| V | 6445 hCoV-19/Japan/PG-1392/2020\|EPI_ISL_684999\|2020-04 | ···················· | ···················· | ····················· | ···················· | ···················· | ···················· |
| V | 6446 hCoV-19/Japan/PG-8171/2020\|EPI_ISL_687270\|2020-07 | ···················· | ···················· | ····················· | ···················· | ···················· | ···················· |
| V | 6447 hCoV-19/Japan/PG-8172/2020\|EPI_ISL_687271\|2020-07 | ···················· | ···················· | ····················· | ···················· | ···················· | ···················· |
| V | 6448 hCoV-19/Japan/PG-8173/2020\|EPI_ISL_687272\|2020-07 | ···················· | ···················· | ····················· | ···················· | ···················· | ···················· |
| V | 6449 hCoV-19/Japan/PG-8174/2020\|EPI_ISL_687273\|2020-07 | ···················· | ···················· | ····················· | ···················· | ···················· | ···················· |
| V | 6450 hCoV-19/Japan/PG-8175/2020\|EPI_ISL_687274\|2020-07 | ···················· | ···················· | ····················· | ···················· | ···················· | ···················· |
| V | 6451 hCoV-19/Japan/PG-8176/2020\|EPI_ISL_687275\|2020-07 | ···················· | ···················· | ····················· | ···················· | ···················· | ···················· |

| | | | | | | | |
|---|---|---|---|---|---|---|---|
| V | 6452 hCoV-19/Japan/PG-8177/2020\|EPI_ISL_687276\|2020-08 | .................... | .................... | .................... | .................... | .................... | .................... |
| V | 6453 hCoV-19/Japan/PG-8178/2020\|EPI_ISL_687277\|2020-08 | .................... | .................... | .................... | .................... | .................... | .................... |
| V | 6454 hCoV-19/Japan/PG-8179/2020\|EPI_ISL_687278\|2020-08 | .................... | .................... | .................... | .................... | .................... | .................... |
| V | 6455 hCoV-19/Japan/PG-8180/2020\|EPI_ISL_687279\|2020-08 | .................... | .................... | .................... | .................... | .................... | .................... |
| V | 6456 hCoV-19/Japan/PG-8181/2020\|EPI_ISL_687280\|2020-08 | .................... | .................... | .................... | .................... | .................... | .................... |
| V | 6457 hCoV-19/Japan/PG-8182/2020\|EPI_ISL_687281\|2020-08 | .................... | .................... | .................... | .................... | .................... | .................... |
| V | 6458 hCoV-19/Japan/PG-8183/2020\|EPI_ISL_687282\|2020-08 | .................... | .................... | .................... | .................... | .................... | .................... |
| V | 6459 hCoV-19/Japan/PG-8184/2020\|EPI_ISL_687283\|2020-08 | .................... | .................... | .................... | .................... | .................... | .................... |
| V | 6460 hCoV-19/Japan/PG-8185/2020\|EPI_ISL_687284\|2020-08 | .................... | .................... | .................... | .................... | .................... | .................... |
| V | 6461 hCoV-19/Japan/PG-8186/2020\|EPI_ISL_687285\|2020-08 | .................... | .................... | .................... | .................... | .................... | .................... |
| V | 6462 hCoV-19/Japan/PG-8187/2020\|EPI_ISL_687286\|2020-08 | .................... | .................... | .................... | .................... | .................... | .................... |
| V | 6463 hCoV-19/Japan/PG-8188/2020\|EPI_ISL_687287\|2020-08 | .................... | .................... | .................... | .................... | .................... | .................... |
| V | 6464 hCoV-19/Japan/PG-8189/2020\|EPI_ISL_687288\|2020-08 | .................... | .................... | .................... | .................... | .................... | .................... |
| V | 6465 hCoV-19/Japan/PG-8190/2020\|EPI_ISL_687289\|2020-08 | .................... | .................... | .................... | .................... | .................... | .................... |
| V | 6466 hCoV-19/Japan/PG-8191/2020\|EPI_ISL_687290\|2020-08 | .................... | .................... | .................... | .................... | .................... | .................... |
| V | 6467 hCoV-19/Japan/PG-8192/2020\|EPI_ISL_687291\|2020-08 | .................... | .................... | .................... | .................... | .................... | .................... |
| V | 6468 hCoV-19/Japan/PG-8193/2020\|EPI_ISL_687292\|2020-08 | .................... | .................... | .................... | .................... | .................... | .................... |
| V | 6469 hCoV-19/Japan/PG-8194/2020\|EPI_ISL_687293\|2020-08 | .................... | .................... | .................... | .................... | .................... | .................... |
| V | 6470 hCoV-19/Japan/PG-8195/2020\|EPI_ISL_687294\|2020-08 | .................... | .................... | .................... | .................... | .................... | .................... |
| V | 6471 hCoV-19/Japan/PG-8196/2020\|EPI_ISL_687295\|2020-08 | .................... | .................... | .................... | .................... | .................... | .................... |
| V | 6472 hCoV-19/Japan/PG-8197/2020\|EPI_ISL_687296\|2020-08 | .................... | .................... | .................... | .................... | .................... | .................... |
| V | 6473 hCoV-19/Japan/PG-8198/2020\|EPI_ISL_687297\|2020-08 | .................... | .................... | .................... | .................... | .................... | .................... |
| V | 6474 hCoV-19/Japan/PG-8199/2020\|EPI_ISL_687298\|2020-08 | .................... | .................... | .................... | .................... | .................... | .................... |
| V | 6475 hCoV-19/Japan/PG-8200/2020\|EPI_ISL_687299\|2020-08 | .................... | .................... | .................... | .................... | .................... | .................... |
| V | 6476 hCoV-19/Japan/PG-4663/2020\|EPI_ISL_689800\|2020-08 | .................... | .................... | .................... | .................... | .................... | .................... |
| V | 6477 hCoV-19/Japan/PG-4664/2020\|EPI_ISL_689801\|2020-08 | .................... | .................... | .................... | .................... | .................... | .................... |
| V | 6478 hCoV-19/Japan/PG-4665/2020\|EPI_ISL_689802\|2020-08 | .................... | .................... | .................... | .................... | .................... | .................... |
| V | 6479 hCoV-19/Japan/PG-4666/2020\|EPI_ISL_689803\|2020-08 | .................... | .................... | .................... | .................... | .................... | .................... |
| V | 6480 hCoV-19/Japan/PG-4667/2020\|EPI_ISL_689804\|2020-08 | .................... | .................... | .................... | .................... | .................... | .................... |
| V | 6481 hCoV-19/Japan/PG-4668/2020\|EPI_ISL_689805\|2020-08 | .................... | .................... | .................... | .................... | .................... | .................... |
| V | 6482 hCoV-19/Japan/PG-4669/2020\|EPI_ISL_689806\|2020-08 | .................... | .................... | .................... | .................... | .................... | .................... |
| V | 6483 hCoV-19/Japan/PG-4670/2020\|EPI_ISL_689807\|2020-08 | .................... | .................... | .................... | .................... | .................... | .................... |
| V | 6484 hCoV-19/Japan/PG-4671/2020\|EPI_ISL_689808\|2020-08 | .................... | .................... | .................... | .................... | .................... | .................... |
| V | 6485 hCoV-19/Japan/PG-4672/2020\|EPI_ISL_689809\|2020-08 | .................... | .................... | .................... | .................... | .................... | .................... |
| V | 6486 hCoV-19/Japan/PG-4673/2020\|EPI_ISL_689810\|2020-08 | .................... | .................... | .................... | .................... | .................... | .................... |
| V | 6487 hCoV-19/Japan/PG-4674/2020\|EPI_ISL_689811\|2020-08 | .................... | .................... | .................... | .................... | .................... | .................... |
| V | 6488 hCoV-19/Japan/PG-4675/2020\|EPI_ISL_689812\|2020-08 | .................... | .................... | .................... | .................... | .................... | .................... |
| V | 6489 hCoV-19/Japan/PG-4676/2020\|EPI_ISL_689813\|2020-08 | .................... | .................... | .................... | .................... | .................... | .................... |
| V | 6490 hCoV-19/Japan/PG-4677/2020\|EPI_ISL_689814\|2020-08 | .................... | .................... | .................... | .................... | .................... | .................... |
| V | 6491 hCoV-19/Japan/PG-4678/2020\|EPI_ISL_689815\|2020-08 | .................... | .................... | .................... | .................... | .................... | .................... |
| V | 6492 hCoV-19/Japan/PG-4679/2020\|EPI_ISL_689816\|2020-08 | .................... | .................... | .................... | .................... | .................... | .................... |
| V | 6493 hCoV-19/Japan/PG-4680/2020\|EPI_ISL_689817\|2020-08 | .................... | .................... | .................... | .................... | .................... | .................... |
| V | 6494 hCoV-19/Japan/PG-4681/2020\|EPI_ISL_689818\|2020-08 | .................... | .................... | .................... | .................... | .................... | .................... |
| V | 6495 hCoV-19/Japan/PG-4682/2020\|EPI_ISL_689819\|2020-08 | .................... | .................... | .................... | .................... | .................... | .................... |
| V | 6496 hCoV-19/Japan/PG-4683/2020\|EPI_ISL_689820\|2020-08 | .................... | .................... | .................... | .................... | .................... | .................... |
| V | 6497 hCoV-19/Japan/PG-4684/2020\|EPI_ISL_689821\|2020-08 | .................... | .................... | .................... | .................... | .................... | .................... |
| V | 6498 hCoV-19/Japan/PG-4685/2020\|EPI_ISL_689822\|2020-08 | .................... | .................... | .................... | .................... | .................... | .................... |
| V | 6499 hCoV-19/Japan/PG-4686/2020\|EPI_ISL_689823\|2020-08 | .................... | .................... | .................... | .................... | .................... | .................... |
| V | 6500 hCoV-19/Japan/PG-4687/2020\|EPI_ISL_689824\|2020-08 | .................... | .................... | .................... | .................... | .................... | .................... |
| V | 6501 hCoV-19/Japan/PG-4688/2020\|EPI_ISL_689825\|2020-08 | .................... | .................... | .................... | .................... | .................... | .................... |
| V | 6502 hCoV-19/Japan/PG-4689/2020\|EPI_ISL_689826\|2020-08 | .................... | .................... | .................... | .................... | .................... | .................... |
| V | 6503 hCoV-19/Japan/PG-4690/2020\|EPI_ISL_689827\|2020-08 | .................... | .................... | .................... | .................... | .................... | .................... |
| V | 6504 hCoV-19/Japan/PG-4691/2020\|EPI_ISL_689828\|2020-08 | .................... | .................... | .................... | .................... | .................... | .................... |
| V | 6505 hCoV-19/Japan/PG-4692/2020\|EPI_ISL_689829\|2020-08 | .................... | .................... | .................... | .................... | .................... | .................... |
| V | 6506 hCoV-19/Japan/PG-4693/2020\|EPI_ISL_689830\|2020-08 | .................... | .................... | .................... | .................... | .................... | .................... |
| V | 6507 hCoV-19/Japan/PG-4694/2020\|EPI_ISL_689831\|2020-08 | .................... | .................... | .................... | .................... | .................... | .................... |
| V | 6508 hCoV-19/Japan/PG-4695/2020\|EPI_ISL_689832\|2020-08 | .................... | .................... | .................... | .................... | .................... | .................... |
| V | 6509 hCoV-19/Japan/PG-4696/2020\|EPI_ISL_689833\|2020-08 | .................... | .................... | .................... | .................... | .................... | .................... |
| V | 6510 hCoV-19/Japan/PG-4697/2020\|EPI_ISL_689834\|2020-08 | .................... | .................... | .................... | .................... | .................... | .................... |
| V | 6511 hCoV-19/Japan/PG-4698/2020\|EPI_ISL_689835\|2020-08 | .................... | .................... | .................... | .................... | .................... | .................... |
| V | 6512 hCoV-19/Japan/PG-4699/2020\|EPI_ISL_689836\|2020-08 | .................... | .................... | .................... | .................... | .................... | .................... |
| V | 6513 hCoV-19/Japan/PG-4700/2020\|EPI_ISL_689837\|2020-07 | .................... | .................... | .................... | .................... | .................... | .................... |
| V | 6514 hCoV-19/Japan/PG-4701/2020\|EPI_ISL_689838\|2020-07 | .................... | .................... | .................... | .................... | .................... | .................... |
| V | 6515 hCoV-19/Japan/PG-4702/2020\|EPI_ISL_689839\|2020-07 | .................... | .................... | .................... | .................... | .................... | .................... |
| V | 6516 hCoV-19/Japan/PG-4703/2020\|EPI_ISL_689840\|2020-08 | .................... | .................... | .................... | .................... | .................... | .................... |
| V | 6517 hCoV-19/Japan/PG-4734/2020\|EPI_ISL_689870\|2020-07 | .................... | .................... | .................... | .................... | .................... | .................... |
| V | 6518 hCoV-19/Japan/PG-4735/2020\|EPI_ISL_689871\|2020-07 | .................... | .................... | .................... | .................... | .................... | .................... |
| V | 6519 hCoV-19/Japan/PG-4736/2020\|EPI_ISL_689872\|2020-07 | .................... | .................... | .................... | .................... | .................... | .................... |
| V | 6520 hCoV-19/Japan/PG-4737/2020\|EPI_ISL_689873\|2020-07 | .................... | .................... | .................... | .................... | .................... | .................... |
| V | 6521 hCoV-19/Japan/PG-4738/2020\|EPI_ISL_689874\|2020-07 | .................... | .................... | .................... | .................... | .................... | .................... |
| V | 6522 hCoV-19/Japan/PG-4739/2020\|EPI_ISL_689875\|2020-07 | .................... | .................... | .................... | .................... | .................... | .................... |
| V | 6523 hCoV-19/Japan/PG-4740/2020\|EPI_ISL_689876\|2020-07 | .................... | .................... | .................... | .................... | .................... | .................... |
| V | 6524 hCoV-19/Japan/PG-4741/2020\|EPI_ISL_689877\|2020-08 | .................... | .................... | .................... | .................... | .................... | .................... |
| V | 6525 hCoV-19/Japan/PG-4742/2020\|EPI_ISL_689878\|2020-08 | .................... | .................... | .................... | .................... | .................... | .................... |
| V | 6526 hCoV-19/Japan/PG-4743/2020\|EPI_ISL_689879\|2020-08 | .................... | .................... | .................... | .................... | .................... | .................... |
| V | 6527 hCoV-19/Japan/PG-4744/2020\|EPI_ISL_689880\|2020-08 | .................... | .................... | .................... | .................... | .................... | .................... |
| V | 6528 hCoV-19/Japan/PG-4745/2020\|EPI_ISL_689881\|2020-08 | .................... | .................... | .................... | .................... | .................... | .................... |
| V | 6529 hCoV-19/Japan/PG-4746/2020\|EPI_ISL_689882\|2020-07 | .................... | .................... | .................... | .................... | .................... | .................... |
| V | 6530 hCoV-19/Japan/PG-4747/2020\|EPI_ISL_689883\|2020-07 | .................... | .................... | .................... | .................... | .................... | .................... |
| V | 6531 hCoV-19/Japan/PG-4748/2020\|EPI_ISL_689884\|2020-07 | .................... | .................... | .................... | .................... | .................... | .................... |

| | | | | | | | |
|---|---|---|---|---|---|---|---|
| V | 6532 hCoV-19/Japan/PG-4749/2020\|EPI_ISL_689885\|2020-07 | .................... | .................... | ..................... | .................... | .................... | .................... |
| V | 6533 hCoV-19/Japan/PG-4750/2020\|EPI_ISL_689886\|2020-07 | .................... | .................... | ..................... | .................... | .................... | .................... |
| V | 6534 hCoV-19/Japan/PG-4751/2020\|EPI_ISL_689887\|2020-07 | .................... | .................... | ..................... | .................... | .................... | .................... |
| V | 6535 hCoV-19/Japan/PG-4752/2020\|EPI_ISL_689888\|2020-08 | .................... | .................... | ..................... | .................... | .................... | .................... |
| V | 6536 hCoV-19/Japan/PG-4753/2020\|EPI_ISL_689889\|2020-08 | .................... | .................... | ..................... | .................... | .................... | .................... |
| V | 6537 hCoV-19/Japan/PG-4754/2020\|EPI_ISL_689890\|2020-08 | .................... | .................... | ..................... | .................... | .................... | .................... |
| V | 6538 hCoV-19/Japan/PG-4755/2020\|EPI_ISL_689891\|2020-08 | .................... | .................... | ..................... | .................... | .................... | .................... |
| V | 6539 hCoV-19/Japan/PG-4756/2020\|EPI_ISL_689892\|2020-08 | .................... | .................... | ..................... | .................... | .................... | .................... |
| V | 6540 hCoV-19/Japan/PG-4757/2020\|EPI_ISL_689893\|2020-08 | .................... | .................... | ..................... | .................... | .................... | .................... |
| V | 6541 hCoV-19/Japan/PG-4758/2020\|EPI_ISL_689894\|2020-08 | .................... | .................... | ..................... | .................... | .................... | .................... |
| V | 6542 hCoV-19/Japan/PG-4759/2020\|EPI_ISL_689895\|2020-08 | .................... | .................... | ..................... | .................... | .................... | .................... |
| V | 6543 hCoV-19/Japan/PG-4760/2020\|EPI_ISL_689896\|2020-08 | .................... | .................... | ..................... | .................... | .................... | .................... |
| V | 6544 hCoV-19/Japan/PG-4761/2020\|EPI_ISL_689897\|2020-08 | .................... | .................... | ..................... | .................... | .................... | .................... |
| V | 6545 hCoV-19/Japan/PG-4762/2020\|EPI_ISL_689898\|2020-08 | .................... | .................... | ..................... | .................... | .................... | .................... |
| V | 6546 hCoV-19/Japan/PG-4763/2020\|EPI_ISL_689899\|2020-08 | .................... | .................... | ..................... | .................... | .................... | .................... |
| V | 6547 hCoV-19/Japan/PG-2668/2020\|EPI_ISL_690800\|2020-06 | .................... | .................... | ..................... | .................... | .................... | .................... |
| V | 6548 hCoV-19/Japan/PG-2669/2020\|EPI_ISL_690801\|2020-06 | .................... | .................... | ..................... | .................... | .................... | .................... |
| V | 6549 hCoV-19/Japan/PG-2670/2020\|EPI_ISL_690802\|2020-06 | .................... | .................... | ..................... | .................... | .................... | .................... |
| V | 6550 hCoV-19/Japan/PG-2671/2020\|EPI_ISL_690803\|2020-06 | .................... | .................... | ..................... | .................... | .................... | .................... |
| V | 6551 hCoV-19/Japan/PG-2672/2020\|EPI_ISL_690804\|2020-06 | .................... | .................... | ..................... | .................... | .................... | .................... |
| V | 6552 hCoV-19/Japan/PG-2673/2020\|EPI_ISL_690805\|2020-06 | .................... | .................... | ..................... | .................... | .................... | .................... |
| V | 6553 hCoV-19/Japan/PG-2674/2020\|EPI_ISL_690806\|2020-06 | .................... | .................... | ..................... | .................... | .................... | .................... |
| V | 6554 hCoV-19/Japan/PG-2675/2020\|EPI_ISL_690807\|2020-06 | .................... | .................... | ..................... | .................... | .................... | .................... |
| V | 6555 hCoV-19/Japan/PG-2676/2020\|EPI_ISL_690808\|2020-06 | .................... | .................... | ..................... | .................... | .................... | .................... |
| V | 6556 hCoV-19/Japan/PG-2677/2020\|EPI_ISL_690809\|2020-06 | .................... | .................... | ..................... | .................... | .................... | .................... |
| V | 6557 hCoV-19/Japan/PG-2678/2020\|EPI_ISL_690810\|2020-06 | .................... | .................... | ..................... | .................... | .................... | .................... |
| V | 6558 hCoV-19/Japan/PG-2679/2020\|EPI_ISL_690811\|2020-06 | .................... | .................... | ..................... | .................... | .................... | .................... |
| V | 6559 hCoV-19/Japan/PG-2680/2020\|EPI_ISL_690812\|2020-06 | .................... | .................... | ..................... | .................... | .................... | .................... |
| V | 6560 hCoV-19/Japan/PG-2681/2020\|EPI_ISL_690813\|2020-06 | .................... | .................... | ..................... | .................... | .................... | .................... |
| V | 6561 hCoV-19/Japan/PG-2682/2020\|EPI_ISL_690814\|2020-06 | .................... | .................... | ..................... | .................... | .................... | .................... |
| V | 6562 hCoV-19/Japan/PG-2683/2020\|EPI_ISL_690815\|2020-06 | .................... | .................... | ..................... | .................... | .................... | .................... |
| V | 6563 hCoV-19/Japan/PG-2684/2020\|EPI_ISL_690816\|2020-06 | .................... | .................... | ..................... | .................... | .................... | .................... |
| V | 6564 hCoV-19/Japan/PG-2685/2020\|EPI_ISL_690817\|2020-06 | .................... | .................... | ..................... | .................... | .................... | .................... |
| V | 6565 hCoV-19/Japan/PG-2686/2020\|EPI_ISL_690818\|2020-03 | .................... | .................... | ..................... | .................... | .................... | .................... |
| V | 6566 hCoV-19/Japan/PG-2687/2020\|EPI_ISL_690819\|2020-03 | .................... | .................... | ..................... | .................... | .................... | .................... |
| V | 6567 hCoV-19/Japan/PG-2688/2020\|EPI_ISL_690820\|2020-03 | .................... | .................... | ..................... | .................... | .................... | .................... |
| V | 6568 hCoV-19/Japan/PG-2689/2020\|EPI_ISL_690821\|2020-03 | .................... | .................... | ..................... | .................... | .................... | .................... |
| V | 6569 hCoV-19/Japan/PG-2690/2020\|EPI_ISL_690822\|2020-03 | .................... | .................... | ..................... | .................... | .................... | .................... |
| V | 6570 hCoV-19/Japan/PG-2691/2020\|EPI_ISL_690823\|2020-03 | .................... | .................... | ..................... | .................... | .................... | .................... |
| V | 6571 hCoV-19/Japan/PG-2692/2020\|EPI_ISL_690824\|2020-03 | .................... | .................... | ..................... | .................... | .................... | .................... |
| V | 6572 hCoV-19/Japan/PG-2693/2020\|EPI_ISL_690825\|2020-03 | .................... | .................... | ..................... | .................... | .................... | .................... |
| V | 6573 hCoV-19/Japan/PG-2694/2020\|EPI_ISL_690826\|2020-03 | .................... | .................... | ..................... | .................... | .................... | .................... |
| V | 6574 hCoV-19/Japan/PG-2695/2020\|EPI_ISL_690827\|2020-04 | .................... | .................... | ..................... | .................... | .................... | .................... |
| V | 6575 hCoV-19/Japan/PG-2696/2020\|EPI_ISL_690828\|2020-04 | .................... | .................... | ..................... | .................... | .................... | .................... |
| V | 6576 hCoV-19/Japan/PG-2697/2020\|EPI_ISL_690829\|2020-04 | .................... | .................... | ..................... | .................... | .................... | .................... |
| V | 6577 hCoV-19/Japan/PG-2719/2020\|EPI_ISL_690851\|2020-03 | .................... | .................... | ..................... | .................... | .................... | .................... |
| V | 6578 hCoV-19/Japan/PG-2728/2020\|EPI_ISL_690860\|2020-03 | .................... | .................... | ..................... | .................... | .................... | .................... |
| V | 6579 hCoV-19/Japan/PG-2729/2020\|EPI_ISL_690861\|2020-03 | .................... | .................... | ..................... | .................... | .................... | .................... |
| V | 6580 hCoV-19/Japan/PG-2730/2020\|EPI_ISL_690862\|2020-03 | .................... | .................... | ..................... | .................... | .................... | .................... |
| V | 6581 hCoV-19/Japan/PG-2731/2020\|EPI_ISL_690863\|2020-03 | .................... | .................... | ..................... | .................... | .................... | .................... |
| V | 6582 hCoV-19/Japan/PG-2732/2020\|EPI_ISL_690864\|2020-03 | .................... | .................... | ..................... | .................... | .................... | .................... |
| V | 6583 hCoV-19/Japan/PG-2733/2020\|EPI_ISL_690865\|2020-03 | .................... | .................... | ..................... | .................... | .................... | .................... |
| V | 6584 hCoV-19/Japan/PG-2734/2020\|EPI_ISL_690866\|2020-03 | .................... | .................... | ..................... | .................... | .................... | .................... |
| V | 6585 hCoV-19/Japan/PG-2735/2020\|EPI_ISL_690867\|2020-03 | .................... | .................... | ..................... | .................... | .................... | .................... |
| V | 6586 hCoV-19/Japan/PG-2736/2020\|EPI_ISL_690868\|2020-03 | .................... | .................... | ..................... | .................... | .................... | .................... |
| V | 6587 hCoV-19/Japan/PG-2737/2020\|EPI_ISL_690869\|2020-03 | .................... | .................... | ..................... | .................... | .................... | .................... |
| V | 6588 hCoV-19/Japan/PG-2738/2020\|EPI_ISL_690870\|2020-03 | .................... | .................... | ..................... | .................... | .................... | .................... |
| V | 6589 hCoV-19/Japan/PG-2739/2020\|EPI_ISL_690871\|2020-03 | .................... | .................... | ..................... | .................... | .................... | .................... |
| V | 6590 hCoV-19/Japan/PG-2740/2020\|EPI_ISL_690872\|2020-03 | .................... | .................... | ..................... | .................... | .................... | .................... |
| V | 6591 hCoV-19/Japan/PG-2741/2020\|EPI_ISL_690873\|2020-03 | .................... | .................... | ..................... | .................... | .................... | .................... |
| V | 6592 hCoV-19/Japan/PG-2742/2020\|EPI_ISL_690874\|2020-04 | .................... | .................... | ..................... | .................... | .................... | .................... |
| V | 6593 hCoV-19/Japan/PG-2743/2020\|EPI_ISL_690875\|2020-04 | .................... | .................... | ..................... | .................... | .................... | .................... |
| V | 6594 hCoV-19/Japan/PG-2744/2020\|EPI_ISL_690876\|2020-04 | .................... | .................... | ..................... | .................... | .................... | .................... |
| V | 6595 hCoV-19/Japan/PG-2745/2020\|EPI_ISL_690877\|2020-04 | .................... | .................... | ..................... | .................... | .................... | .................... |
| V | 6596 hCoV-19/Japan/PG-2746/2020\|EPI_ISL_690878\|2020-04 | .................... | .................... | ..................... | .................... | .................... | .................... |
| V | 6597 hCoV-19/Japan/PG-2747/2020\|EPI_ISL_690879\|2020-04 | .................... | .................... | ..................... | .................... | .................... | .................... |
| V | 6598 hCoV-19/Japan/PG-2748/2020\|EPI_ISL_690880\|2020-04 | .................... | .................... | ..................... | .................... | .................... | .................... |
| V | 6599 hCoV-19/Japan/PG-2749/2020\|EPI_ISL_690881\|2020-04 | .................... | .................... | ..................... | .................... | .................... | .................... |
| V | 6600 hCoV-19/Japan/PG-2750/2020\|EPI_ISL_690882\|2020-04 | .................... | .................... | ..................... | .................... | .................... | .................... |
| V | 6601 hCoV-19/Japan/PG-2751/2020\|EPI_ISL_690883\|2020-04 | .................... | .................... | ..................... | .................... | .................... | .................... |
| V | 6602 hCoV-19/Japan/PG-2752/2020\|EPI_ISL_690884\|2020-04 | .................... | .................... | ..................... | .................... | .................... | .................... |
| V | 6603 hCoV-19/Japan/PG-2753/2020\|EPI_ISL_690885\|2020-04 | .................... | .................... | ..................... | .................... | .................... | .................... |
| V | 6604 hCoV-19/Japan/PG-2754/2020\|EPI_ISL_690886\|2020-04 | .................... | .................... | ..................... | .................... | .................... | .................... |
| V | 6605 hCoV-19/Japan/PG-2755/2020\|EPI_ISL_690887\|2020-04 | .................... | .................... | ..................... | .................... | .................... | .................... |
| V | 6606 hCoV-19/Japan/PG-2756/2020\|EPI_ISL_690888\|2020-04 | .................... | .................... | ..................... | .................... | .................... | .................... |
| V | 6607 hCoV-19/Japan/PG-2757/2020\|EPI_ISL_690889\|2020-04 | .................... | .................... | ..................... | .................... | .................... | .................... |
| V | 6608 hCoV-19/Japan/PG-2758/2020\|EPI_ISL_690890\|2020-04 | .................... | .................... | ..................... | .................... | .................... | .................... |
| V | 6609 hCoV-19/Japan/PG-2759/2020\|EPI_ISL_690891\|2020-04 | .................... | .................... | ..................... | .................... | .................... | .................... |
| V | 6610 hCoV-19/Japan/PG-2760/2020\|EPI_ISL_690892\|2020-04 | .................... | .................... | ..................... | .................... | .................... | .................... |
| V | 6611 hCoV-19/Japan/PG-2761/2020\|EPI_ISL_690893\|2020-04 | .................... | .................... | ..................... | .................... | .................... | .................... |

| | | | | | | | |
|---|---|---|---|---|---|---|---|
| V | 6612 hCoV-19/Japan/PG-2762/2020\|EPI_ISL_690894\|2020-04 | ···················· | ···················· | ····················· | ···················· | ···················· | ···················· |
| V | 6613 hCoV-19/Japan/PG-2763/2020\|EPI_ISL_690895\|2020-04 | ···················· | ·A·················· | ····················· | ···················· | ···················· | ···················· |
| V | 6614 hCoV-19/Japan/PG-2764/2020\|EPI_ISL_690896\|2020-04 | ···················· | ···················· | ····················· | ···················· | ···················· | ···················· |
| V | 6615 hCoV-19/Japan/PG-2765/2020\|EPI_ISL_690897\|2020-04 | ···················· | ···················· | ····················· | ···················· | ···················· | ···················· |
| V | 6616 hCoV-19/Japan/PG-2766/2020\|EPI_ISL_690898\|2020-04 | ···················· | ···················· | ····················· | ···················· | ···················· | ···················· |
| V | 6617 hCoV-19/Japan/PG-2767/2020\|EPI_ISL_690899\|2020-04 | ···················· | ···················· | ····················· | ···················· | ···················· | ···················· |
| V | 6618 hCoV-19/USA/MA-MASPHL-00559/2020\|EPI_ISL_693028\|20 | ···················· | ···················· | ····················· | ···················· | ···················· | ···················· |
| V | 6619 hCoV-19/USA/MA-MASPHL-00557/2020\|EPI_ISL_693030\|20 | ···················· | ···················· | ····················· | ···················· | ···················· | ···················· |
| V | 6620 hCoV-19/USA/MA-MASPHL-00555/2020\|EPI_ISL_693032\|20 | ···················· | ···················· | ····················· | ···················· | ···················· | ···················· |
| V | 6621 hCoV-19/USA/MA-MASPHL-00550/2020\|EPI_ISL_693036\|20 | ···················· | ···················· | ····················· | ···················· | ···················· | ···················· |
| V | 6622 hCoV-19/USA/MA-MASPHL-00549/2020\|EPI_ISL_693037\|20 | ···················· | ···················· | ····················· | ···················· | ···················· | ···················· |
| V | 6623 hCoV-19/USA/MA-MASPHL-00545/2020\|EPI_ISL_693039\|20 | ···················· | ···················· | ····················· | ···················· | ···················· | ···················· |
| V | 6624 hCoV-19/USA/MA-MASPHL-00536/2020\|EPI_ISL_693043\|20 | ···················· | ···················· | ····················· | ···················· | ···················· | ···················· |
| V | 6625 hCoV-19/USA/MA-MASPHL-00534/2020\|EPI_ISL_693045\|20 | ···················· | ···················· | ····················· | ···················· | ···················· | ···················· |
| V | 6626 hCoV-19/USA/MA-MASPHL-00533/2020\|EPI_ISL_693046\|20 | ···················· | ···················· | ····················· | ···················· | ···················· | ···················· |
| V | 6627 hCoV-19/USA/MA-MASPHL-00531/2020\|EPI_ISL_693047\|20 | ···················· | ···················· | ····················· | ···················· | ···················· | ···················· |
| V | 6628 hCoV-19/USA/MA-MASPHL-00530/2020\|EPI_ISL_693048\|20 | ···················· | ···················· | ····················· | ···················· | ···················· | ···················· |
| V | 6629 hCoV-19/USA/MA-MASPHL-00529/2020\|EPI_ISL_693049\|20 | ···················· | ···················· | ····················· | ···················· | ···················· | ···················· |
| V | 6630 hCoV-19/USA/MA-MASPHL-00527/2020\|EPI_ISL_693051\|20 | ···················· | ···················· | ····················· | ···················· | ···················· | ···················· |
| V | 6631 hCoV-19/USA/MA-MASPHL-00526/2020\|EPI_ISL_693052\|20 | ···················· | ···················· | ····················· | ···················· | ···················· | ···················· |
| V | 6632 hCoV-19/USA/MA-MASPHL-00519/2020\|EPI_ISL_693058\|20 | ···················· | ···················· | ····················· | ···················· | ···················· | ···················· |
| V | 6633 hCoV-19/USA/MA-MASPHL-00517/2020\|EPI_ISL_693060\|20 | ···················· | ···················· | ····················· | ···················· | ···················· | ···················· |
| V | 6634 hCoV-19/USA/MA-MASPHL-00512/2020\|EPI_ISL_693062\|20 | ···················· | ···················· | ····················· | ···················· | ···················· | ···················· |
| V | 6635 hCoV-19/USA/MA-MASPHL-00511/2020\|EPI_ISL_693063\|20 | ···················· | ···················· | ····················· | ···················· | ···················· | ···················· |
| V | 6636 hCoV-19/USA/MA-MASPHL-00510/2020\|EPI_ISL_693064\|20 | ···················· | ···················· | ····················· | ···················· | ···················· | ···················· |
| V | 6637 hCoV-19/USA/MA-MASPHL-00509/2020\|EPI_ISL_693065\|20 | ···················· | ···················· | ····················· | ···················· | ···················· | ···················· |
| V | 6638 hCoV-19/USA/MA-MASPHL-00506/2020\|EPI_ISL_693066\|20 | ···················· | ···················· | ····················· | ···················· | ···················· | ···················· |
| V | 6639 hCoV-19/USA/MA-MASPHL-00504/2020\|EPI_ISL_693067\|20 | ···················· | ···················· | ····················· | ···················· | ···················· | ···················· |
| V | 6640 hCoV-19/USA/MA-MASPHL-00501/2020\|EPI_ISL_693069\|20 | ···················· | ···················· | ····················· | ···················· | ···················· | ···················· |
| V | 6641 hCoV-19/USA/MA-MASPHL-00500/2020\|EPI_ISL_693070\|20 | ···················· | ···················· | ····················· | ···················· | ···················· | ···················· |
| V | 6642 hCoV-19/USA/MA-MASPHL-00499/2020\|EPI_ISL_693071\|20 | ···················· | ···················· | ····················· | ···················· | ···················· | ···················· |
| V | 6643 hCoV-19/USA/MA-MASPHL-00498/2020\|EPI_ISL_693072\|20 | ···················· | ···················· | ····················· | ···················· | ···················· | ···················· |
| V | 6644 hCoV-19/USA/MA-MASPHL-00496/2020\|EPI_ISL_693074\|20 | ···················· | ···················· | ····················· | ···················· | ···················· | ···················· |
| V | 6645 hCoV-19/USA/MA-MASPHL-00495/2020\|EPI_ISL_693075\|20 | ···················· | ···················· | ····················· | ···················· | ···················· | ···················· |
| V | 6646 hCoV-19/USA/MA-MASPHL-00492/2020\|EPI_ISL_693076\|20 | ···················· | ···················· | ····················· | ···················· | ···················· | ···················· |
| V | 6647 hCoV-19/USA/MA-MASPHL-00490/2020\|EPI_ISL_693077\|20 | ···················· | ···················· | ····················· | ···················· | ···················· | ···················· |
| V | 6648 hCoV-19/USA/MA-MASPHL-00489/2020\|EPI_ISL_693078\|20 | ···················· | ···················· | ····················· | ···················· | ···················· | ···················· |
| V | 6649 hCoV-19/USA/MA-MASPHL-00487/2020\|EPI_ISL_693080\|20 | ···················· | ···················· | ····················· | ···················· | ···················· | ···················· |
| V | 6650 hCoV-19/USA/MA-MASPHL-00485/2020\|EPI_ISL_693082\|20 | ···················· | ···················· | ····················· | ···················· | ···················· | ···················· |
| V | 6651 hCoV-19/USA/MA-MASPHL-00484/2020\|EPI_ISL_693083\|20 | ···················· | ···················· | ····················· | ···················· | ···················· | ···················· |
| V | 6652 hCoV-19/USA/MA-MASPHL-00483/2020\|EPI_ISL_693084\|20 | ···················· | ···················· | ····················· | ···················· | ···················· | ···················· |
| V | 6653 hCoV-19/USA/MA-MASPHL-00475/2020\|EPI_ISL_693090\|20 | ···················· | ···················· | ····················· | ···················· | ···················· | ···················· |
| V | 6654 hCoV-19/USA/MA-MASPHL-00474/2020\|EPI_ISL_693091\|20 | ···················· | ···················· | ····················· | ···················· | ···················· | ···················· |
| V | 6655 hCoV-19/USA/MA-MASPHL-00465/2020\|EPI_ISL_693098\|20 | ···················· | ···················· | ····················· | ···················· | ···················· | ···················· |
| V | 6656 hCoV-19/USA/MA-MASPHL-00463/2020\|EPI_ISL_693099\|20 | ···················· | ···················· | ····················· | ···················· | ···················· | ···················· |
| V | 6657 hCoV-19/Japan/PG-1314/2020\|EPI_ISL_684922\|2020-04 | ···················· | ···················· | ····················· | ···················· | ···················· | ···················· |
| V | 6658 hCoV-19/Japan/PG-1317/2020\|EPI_ISL_684925\|2020-04 | ···················· | ···················· | ····················· | ···················· | ···················· | ···················· |
| V | 6659 hCoV-19/Japan/PG-1318/2020\|EPI_ISL_684926\|2020-04 | ···················· | ···················· | ····················· | ···················· | ···················· | ···················· |
| V | 6660 hCoV-19/Japan/PG-1319/2020\|EPI_ISL_684927\|2020-04 | ···················· | ···················· | ····················· | ···················· | ···················· | ···················· |
| V | 6661 hCoV-19/Japan/PG-1320/2020\|EPI_ISL_684928\|2020-04 | ···················· | ···················· | ····················· | ···················· | ···················· | ···················· |
| V | 6662 hCoV-19/Japan/PG-1321/2020\|EPI_ISL_684929\|2020-04 | ···················· | ···················· | ····················· | ···················· | ···················· | ···················· |
| V | 6663 hCoV-19/Japan/PG-1322/2020\|EPI_ISL_684930\|2020-04 | ···················· | ···················· | ····················· | ···················· | ···················· | ···················· |
| V | 6664 hCoV-19/Japan/PG-1323/2020\|EPI_ISL_684931\|2020-04 | ···················· | ···················· | ····················· | ···················· | ···················· | ···················· |
| V | 6665 hCoV-19/Japan/PG-1324/2020\|EPI_ISL_684932\|2020-04 | ···················· | ···················· | ····················· | ···················· | ···················· | ···················· |
| V | 6666 hCoV-19/Japan/PG-1325/2020\|EPI_ISL_684933\|2020-04 | ···················· | ···················· | ····················· | ···················· | ···················· | ···················· |
| V | 6667 hCoV-19/Japan/PG-1326/2020\|EPI_ISL_684934\|2020-04 | ···················· | ···················· | ····················· | ···················· | ···················· | ···················· |
| V | 6668 hCoV-19/Japan/PG-1327/2020\|EPI_ISL_684935\|2020-04 | ···················· | ···················· | ····················· | ···················· | ···················· | ···················· |
| V | 6669 hCoV-19/Japan/PG-1328/2020\|EPI_ISL_684936\|2020-04 | ···················· | ···················· | ····················· | ···················· | ···················· | ···················· |
| V | 6670 hCoV-19/Japan/PG-1329/2020\|EPI_ISL_684937\|2020-04 | ···················· | ···················· | ····················· | ···················· | ···················· | ···················· |
| V | 6671 hCoV-19/Japan/PG-1330/2020\|EPI_ISL_684938\|2020-04 | ···················· | ···················· | ····················· | ···················· | ···················· | ···················· |
| V | 6672 hCoV-19/Japan/PG-1331/2020\|EPI_ISL_684939\|2020-04 | ···················· | ···················· | ····················· | ···················· | ···················· | ···················· |
| V | 6673 hCoV-19/Japan/PG-1332/2020\|EPI_ISL_684940\|2020-04 | ···················· | ···················· | ····················· | ···················· | ···················· | ···················· |
| V | 6674 hCoV-19/Japan/PG-1333/2020\|EPI_ISL_684941\|2020-04 | ···················· | ···················· | ····················· | ···················· | ···················· | ···················· |
| V | 6675 hCoV-19/Japan/PG-1334/2020\|EPI_ISL_684942\|2020-04 | ···················· | ···················· | ····················· | ···················· | ···················· | ···················· |
| V | 6676 hCoV-19/Japan/PG-1335/2020\|EPI_ISL_684943\|2020-04 | ···················· | ···················· | ····················· | ···················· | ···················· | ···················· |
| V | 6677 hCoV-19/Japan/PG-1336/2020\|EPI_ISL_684944\|2020-04 | ···················· | ···················· | ····················· | ···················· | ···················· | ···················· |
| V | 6678 hCoV-19/Japan/PG-1337/2020\|EPI_ISL_684945\|2020-04 | ···················· | ···················· | ····················· | ···················· | ···················· | ···················· |
| V | 6679 hCoV-19/Japan/PG-1338/2020\|EPI_ISL_684946\|2020-04 | ···················· | ···················· | ····················· | ···················· | ···················· | ···················· |
| V | 6680 hCoV-19/Japan/PG-1339/2020\|EPI_ISL_684947\|2020-04 | ···················· | ···················· | ····················· | ···················· | ···················· | ···················· |
| V | 6681 hCoV-19/Japan/PG-1340/2020\|EPI_ISL_684948\|2020-04 | ···················· | ···················· | ····················· | ···················· | ···················· | ···················· |
| V | 6682 hCoV-19/Japan/PG-1341/2020\|EPI_ISL_684949\|2020-04 | ···················· | ···················· | ····················· | ···················· | ···················· | ···················· |
| V | 6683 hCoV-19/Japan/PG-1342/2020\|EPI_ISL_684950\|2020-04 | ···················· | ···················· | ····················· | ···················· | ···················· | ···················· |
| V | 6684 hCoV-19/Japan/PG-1343/2020\|EPI_ISL_684951\|2020-04 | ···················· | ···················· | ····················· | ···················· | ···················· | ···················· |
| V | 6685 hCoV-19/Japan/PG-1344/2020\|EPI_ISL_684952\|2020-04 | ···················· | ···················· | ····················· | ···················· | ···················· | ···················· |
| V | 6686 hCoV-19/Japan/PG-1345/2020\|EPI_ISL_684953\|2020-04 | ···················· | ···················· | ····················· | ···················· | ···················· | ···················· |
| V | 6687 hCoV-19/Japan/PG-7999/2020\|EPI_ISL_687100\|2020-08 | ···················· | ···················· | ····················· | ···················· | ···················· | ···················· |
| V | 6688 hCoV-19/Japan/PG-8000/2020\|EPI_ISL_687101\|2020-08 | ···················· | ···················· | ····················· | ···················· | ···················· | ···················· |
| V | 6689 hCoV-19/Japan/PG-8001/2020\|EPI_ISL_687102\|2020-08 | ···················· | ···················· | ····················· | ···················· | ···················· | ···················· |
| V | 6690 hCoV-19/Japan/PG-8002/2020\|EPI_ISL_687103\|2020-08 | ···················· | ···················· | ····················· | ···················· | ···················· | ···················· |
| V | 6691 hCoV-19/Japan/PG-8003/2020\|EPI_ISL_687104\|2020-08 | ···················· | ···················· | ····················· | ···················· | ···················· | ···················· |

| | | | | | | | | |
|---|---|---|---|---|---|---|---|---|
| V | 6692 | hCoV-19/Japan/PG-8004/2020\|EPI_ISL_687105\|2020-08 | ···················· | ···················· | ····················· | ···················· | ···················· | ···················· |
| V | 6693 | hCoV-19/Japan/PG-8005/2020\|EPI_ISL_687106\|2020-08 | ···················· | ···················· | ····················· | ···················· | ···················· | ···················· |
| V | 6694 | hCoV-19/Japan/PG-8006/2020\|EPI_ISL_687107\|2020-08 | ···················· | ···················· | ····················· | ···················· | ···················· | ···················· |
| V | 6695 | hCoV-19/Japan/PG-8007/2020\|EPI_ISL_687108\|2020-08 | ···················· | ···················· | ····················· | ···················· | ···················· | ···················· |
| V | 6696 | hCoV-19/Japan/PG-8008/2020\|EPI_ISL_687109\|2020-08 | ···················· | ···················· | ····················· | ···················· | ···················· | ···················· |
| V | 6697 | hCoV-19/Japan/PG-8009/2020\|EPI_ISL_687110\|2020-08 | ···················· | ···················· | ····················· | ···················· | ···················· | ···················· |
| V | 6698 | hCoV-19/Japan/PG-8010/2020\|EPI_ISL_687111\|2020-08 | ···················· | ···················· | ····················· | ···················· | ···················· | ···················· |
| V | 6699 | hCoV-19/Japan/PG-8011/2020\|EPI_ISL_687112\|2020-08 | ···················· | ···················· | ····················· | ···················· | ···················· | ···················· |
| V | 6700 | hCoV-19/Japan/PG-8012/2020\|EPI_ISL_687113\|2020-08 | ···················· | ···················· | ····················· | ···················· | ···················· | ···················· |
| V | 6701 | hCoV-19/Japan/PG-8013/2020\|EPI_ISL_687114\|2020-08 | ···················· | ···················· | ····················· | ···················· | ···················· | ···················· |
| V | 6702 | hCoV-19/Japan/PG-8014/2020\|EPI_ISL_687115\|2020-08 | ···················· | ···················· | ····················· | ···················· | ···················· | ···················· |
| V | 6703 | hCoV-19/Japan/PG-8015/2020\|EPI_ISL_687116\|2020-08 | ···················· | ···················· | ····················· | ···················· | ···················· | ···················· |
| V | 6704 | hCoV-19/Japan/PG-8016/2020\|EPI_ISL_687117\|2020-08 | ···················· | ···················· | ····················· | ···················· | ···················· | ···················· |
| V | 6705 | hCoV-19/Japan/PG-8017/2020\|EPI_ISL_687118\|2020-08 | ···················· | ···················· | ····················· | ···················· | ···················· | ···················· |
| V | 6706 | hCoV-19/Japan/PG-8018/2020\|EPI_ISL_687119\|2020-08 | ···················· | ···················· | ····················· | ···················· | ···················· | ···················· |
| V | 6707 | hCoV-19/Japan/PG-8019/2020\|EPI_ISL_687120\|2020-08 | ···················· | ···················· | ····················· | ···················· | ···················· | ···················· |
| V | 6708 | hCoV-19/Japan/PG-8020/2020\|EPI_ISL_687121\|2020-08 | ···················· | ···················· | ····················· | ···················· | ···················· | ···················· |
| V | 6709 | hCoV-19/Japan/PG-8021/2020\|EPI_ISL_687122\|2020-08 | ···················· | ···················· | ····················· | ···················· | ···················· | ···················· |
| V | 6710 | hCoV-19/Japan/PG-8022/2020\|EPI_ISL_687123\|2020-08 | ···················· | ···················· | ····················· | ···················· | ···················· | ···················· |
| V | 6711 | hCoV-19/Japan/PG-8023/2020\|EPI_ISL_687124\|2020-08 | ···················· | ···················· | ····················· | ···················· | ···················· | ···················· |
| V | 6712 | hCoV-19/Japan/PG-8024/2020\|EPI_ISL_687125\|2020-08 | ···················· | ···················· | ····················· | ···················· | ···················· | ···················· |
| V | 6713 | hCoV-19/Japan/PG-8025/2020\|EPI_ISL_687126\|2020-08 | ···················· | ···················· | ····················· | ···················· | ···················· | ···················· |
| V | 6714 | hCoV-19/Japan/PG-8026/2020\|EPI_ISL_687127\|2020-08 | ···················· | ···················· | ····················· | ···················· | ···················· | ···················· |
| V | 6715 | hCoV-19/Japan/PG-8027/2020\|EPI_ISL_687128\|2020-08 | ···················· | ···················· | ····················· | ···················· | ···················· | ···················· |
| V | 6716 | hCoV-19/Japan/PG-8028/2020\|EPI_ISL_687129\|2020-08 | ···················· | ···················· | ····················· | ···················· | ···················· | ···················· |
| V | 6717 | hCoV-19/Japan/PG-8029/2020\|EPI_ISL_687130\|2020-08 | ···················· | ···················· | ····················· | ···················· | ···················· | ···················· |
| V | 6718 | hCoV-19/Japan/PG-8030/2020\|EPI_ISL_687131\|2020-08 | ···················· | ···················· | ····················· | ···················· | ···················· | ···················· |
| V | 6719 | hCoV-19/Japan/PG-8031/2020\|EPI_ISL_687132\|2020-08 | ···················· | ···················· | ····················· | ···················· | ···················· | ···················· |
| V | 6720 | hCoV-19/Japan/PG-8032/2020\|EPI_ISL_687133\|2020-08 | ···················· | ···················· | ····················· | ···················· | ···················· | ···················· |
| V | 6721 | hCoV-19/Japan/PG-8033/2020\|EPI_ISL_687134\|2020-08 | ········T··········· | ···················· | ····················· | ···················· | ···················· | ···················· |
| V | 6722 | hCoV-19/Japan/PG-8034/2020\|EPI_ISL_687135\|2020-08 | ···················· | ···················· | ····················· | ···················· | ···················· | ···················· |
| V | 6723 | hCoV-19/Japan/PG-8035/2020\|EPI_ISL_687136\|2020-08 | ···················· | ···················· | ····················· | ···················· | ···················· | ···················· |
| V | 6724 | hCoV-19/Japan/PG-8036/2020\|EPI_ISL_687137\|2020-08 | ········T··········· | ···················· | ····················· | ···················· | ···················· | ···················· |
| V | 6725 | hCoV-19/Japan/PG-8037/2020\|EPI_ISL_687138\|2020-09 | ···················· | ···················· | ····················· | ···················· | ···················· | ···················· |
| V | 6726 | hCoV-19/Japan/PG-8038/2020\|EPI_ISL_687139\|2020-08 | ···················· | ···················· | ····················· | ···················· | ···················· | ···················· |
| V | 6727 | hCoV-19/Japan/PG-8039/2020\|EPI_ISL_687140\|2020-08 | ···················· | ···················· | ····················· | ···················· | ···················· | ···················· |
| V | 6728 | hCoV-19/Japan/PG-8040/2020\|EPI_ISL_687141\|2020-08 | ···················· | ···················· | ····················· | ···················· | ···················· | ···················· |
| V | 6729 | hCoV-19/Japan/PG-8041/2020\|EPI_ISL_687142\|2020-08 | ···················· | ···················· | ····················· | ···················· | ···················· | ···················· |
| V | 6730 | hCoV-19/Japan/PG-8042/2020\|EPI_ISL_687143\|2020-08 | ···················· | ···················· | ····················· | ···················· | ···················· | ···················· |
| V | 6731 | hCoV-19/Japan/PG-8043/2020\|EPI_ISL_687144\|2020-08 | ···················· | ···················· | ····················· | ···················· | ···················· | ···················· |
| V | 6732 | hCoV-19/Japan/PG-8044/2020\|EPI_ISL_687145\|2020-08 | ···················· | ···················· | ····················· | ···················· | ···················· | ···················· |
| V | 6733 | hCoV-19/Japan/PG-8045/2020\|EPI_ISL_687146\|2020-08 | ···················· | ···················· | ····················· | ···················· | ···················· | ···················· |
| V | 6734 | hCoV-19/Japan/PG-8046/2020\|EPI_ISL_687147\|2020-08 | ···················· | ···················· | ····················· | ···················· | ···················· | ···················· |
| V | 6735 | hCoV-19/Japan/PG-8047/2020\|EPI_ISL_687148\|2020-09 | ···················· | ···················· | ····················· | ···················· | ···················· | ···················· |
| V | 6736 | hCoV-19/Japan/PG-8048/2020\|EPI_ISL_687149\|2020-09 | ···················· | ···················· | ····················· | ···················· | ···················· | ···················· |
| V | 6737 | hCoV-19/Japan/PG-7297/2020\|EPI_ISL_688400\|2020-08 | ···················· | ···················· | ····················· | ···················· | ···················· | ···················· |
| V | 6738 | hCoV-19/Japan/PG-7298/2020\|EPI_ISL_688401\|2020-08 | ···················· | ···················· | ····················· | ···················· | ···················· | ···················· |
| V | 6739 | hCoV-19/Japan/PG-7299/2020\|EPI_ISL_688402\|2020-08 | ···················· | ···················· | ····················· | ···················· | ···················· | ···················· |
| V | 6740 | hCoV-19/Japan/PG-7300/2020\|EPI_ISL_688403\|2020-08 | ···················· | ···················· | ····················· | ···················· | ···················· | ···················· |
| V | 6741 | hCoV-19/Japan/PG-7301/2020\|EPI_ISL_688404\|2020-08 | ···················· | ···················· | ····················· | ···················· | ···················· | ···················· |
| V | 6742 | hCoV-19/Japan/PG-7302/2020\|EPI_ISL_688405\|2020-08 | ···················· | ···················· | ····················· | ···················· | ···················· | ···················· |
| V | 6743 | hCoV-19/Japan/PG-7303/2020\|EPI_ISL_688406\|2020-08 | ···················· | ···················· | ····················· | ···················· | ···················· | ···················· |
| V | 6744 | hCoV-19/Japan/PG-7304/2020\|EPI_ISL_688407\|2020-08 | ···················· | ···················· | ····················· | ···················· | ···················· | ···················· |
| V | 6745 | hCoV-19/Japan/PG-7305/2020\|EPI_ISL_688408\|2020-08 | ···················· | ···················· | ····················· | ···················· | ···················· | ···················· |
| V | 6746 | hCoV-19/Japan/PG-7306/2020\|EPI_ISL_688409\|2020-08 | ···················· | ···················· | ····················· | ···················· | ···················· | ···················· |
| V | 6747 | hCoV-19/Japan/PG-7307/2020\|EPI_ISL_688410\|2020-08 | ···················· | ···················· | ····················· | ···················· | ···················· | ···················· |
| V | 6748 | hCoV-19/Japan/PG-7308/2020\|EPI_ISL_688411\|2020-08 | ···················· | ···················· | ····················· | ···················· | ···················· | ···················· |
| V | 6749 | hCoV-19/Japan/PG-7309/2020\|EPI_ISL_688412\|2020-08 | ···················· | ···················· | ····················· | ···················· | ···················· | ···················· |
| V | 6750 | hCoV-19/Japan/PG-7310/2020\|EPI_ISL_688413\|2020-08 | ···················· | ···················· | ····················· | ···················· | ···················· | ···················· |
| V | 6751 | hCoV-19/Japan/PG-7311/2020\|EPI_ISL_688414\|2020-08 | ···················· | ···················· | ····················· | ···················· | ···················· | ···················· |
| V | 6752 | hCoV-19/Japan/PG-7312/2020\|EPI_ISL_688415\|2020-08 | ···················· | ···················· | ····················· | ···················· | ···················· | ···················· |
| V | 6753 | hCoV-19/Japan/PG-7313/2020\|EPI_ISL_688416\|2020-08 | ···················· | ···················· | ····················· | ···················· | ···················· | ···················· |
| V | 6754 | hCoV-19/Japan/PG-7314/2020\|EPI_ISL_688417\|2020-08 | ···················· | ···················· | ····················· | ···················· | ···················· | ···················· |
| V | 6755 | hCoV-19/Japan/PG-7315/2020\|EPI_ISL_688418\|2020-08 | ···················· | ···················· | ····················· | ···················· | ···················· | ···················· |
| V | 6756 | hCoV-19/Japan/PG-7316/2020\|EPI_ISL_688419\|2020-08 | ···················· | ···················· | ····················· | ···················· | ···················· | ···················· |
| V | 6757 | hCoV-19/Japan/PG-7317/2020\|EPI_ISL_688420\|2020-08 | ···················· | ···················· | ····················· | ···················· | ···················· | ···················· |
| V | 6758 | hCoV-19/Japan/PG-7318/2020\|EPI_ISL_688421\|2020-08 | ···················· | ···················· | ····················· | ···················· | ···················· | ···················· |
| V | 6759 | hCoV-19/Japan/PG-7319/2020\|EPI_ISL_688422\|2020-08 | ···················· | ···················· | ····················· | ···················· | ···················· | ···················· |
| V | 6760 | hCoV-19/Japan/PG-7320/2020\|EPI_ISL_688423\|2020-08 | ···················· | ···················· | ····················· | ···················· | ···················· | ···················· |
| V | 6761 | hCoV-19/Japan/PG-7321/2020\|EPI_ISL_688424\|2020-08 | ···················· | ···················· | ····················· | ···················· | ···················· | ···················· |
| V | 6762 | hCoV-19/Japan/PG-7322/2020\|EPI_ISL_688425\|2020-08 | ···················· | ···················· | ····················· | ···················· | ···················· | ···················· |
| V | 6763 | hCoV-19/Japan/PG-7323/2020\|EPI_ISL_688426\|2020-08 | ···················· | ···················· | ····················· | ···················· | ···················· | ···················· |
| V | 6764 | hCoV-19/Japan/PG-7324/2020\|EPI_ISL_688427\|2020-08 | ···················· | ···················· | ····················· | ···················· | ···················· | ···················· |
| V | 6765 | hCoV-19/Japan/PG-7325/2020\|EPI_ISL_688428\|2020-08 | ···················· | ···················· | ····················· | ···················· | ···················· | ···················· |
| V | 6766 | hCoV-19/Japan/PG-7326/2020\|EPI_ISL_688429\|2020-08 | ···················· | ···················· | ····················· | ···················· | ···················· | ···················· |
| V | 6767 | hCoV-19/Japan/PG-7327/2020\|EPI_ISL_688430\|2020-08 | ···················· | ···················· | ····················· | ···················· | ···················· | ···················· |
| V | 6768 | hCoV-19/Japan/PG-7328/2020\|EPI_ISL_688431\|2020-08 | ···················· | ···················· | ····················· | ···················· | ···················· | ···················· |
| V | 6769 | hCoV-19/Japan/PG-7329/2020\|EPI_ISL_688432\|2020-08 | ···················· | ···················· | ····················· | ···················· | ···················· | ···················· |
| V | 6770 | hCoV-19/Japan/PG-7330/2020\|EPI_ISL_688433\|2020-08 | ···················· | ···················· | ····················· | ···················· | ···················· | ···················· |
| V | 6771 | hCoV-19/Japan/PG-7331/2020\|EPI_ISL_688434\|2020-08 | ···················· | ···················· | ····················· | ···················· | ···················· | ···················· |

| | | | | | | | | |
|---|---|---|---|---|---|---|---|---|
| V | 6772 hCoV-19/Japan/PG-7332/2020\|EPI_ISL_688435\|2020-08 | ···················· | ···················· | ···················· | ···················· | ···················· | ···················· | |
| V | 6773 hCoV-19/Japan/PG-7333/2020\|EPI_ISL_688436\|2020-08 | ···················· | ···················· | ···················· | ···················· | ···················· | ···················· | |
| V | 6774 hCoV-19/Japan/PG-7334/2020\|EPI_ISL_688437\|2020-08 | ···················· | ···················· | ···················· | ···················· | ···················· | ···················· | |
| V | 6775 hCoV-19/Japan/PG-7335/2020\|EPI_ISL_688438\|2020-08 | ···················· | ···················· | ···················· | ···················· | ···················· | ···················· | |
| V | 6776 hCoV-19/Japan/PG-7336/2020\|EPI_ISL_688439\|2020-08 | ···················· | ···················· | ···················· | ···················· | ···················· | ···················· | |
| V | 6777 hCoV-19/Japan/PG-7337/2020\|EPI_ISL_688440\|2020-08 | ···················· | ···················· | ···················· | ···················· | ···················· | ···················· | |
| V | 6778 hCoV-19/Japan/PG-7338/2020\|EPI_ISL_688441\|2020-08 | ···················· | ···················· | ···················· | ···················· | ···················· | ···················· | |
| V | 6780 hCoV-19/Japan/PG-7340/2020\|EPI_ISL_688443\|2020-08 | ···················· | ···················· | ···················· | ···················· | ···················· | ···················· | |
| V | 6781 hCoV-19/Japan/PG-7341/2020\|EPI_ISL_688444\|2020-08 | ···················· | ···················· | ···················· | ···················· | ···················· | ···················· | |
| V | 6782 hCoV-19/Japan/PG-7342/2020\|EPI_ISL_688445\|2020-08 | ···················· | ···················· | ···················· | ···················· | ···················· | ···················· | |
| V | 6783 hCoV-19/Japan/PG-7343/2020\|EPI_ISL_688446\|2020-08 | ···················· | ···················· | ···················· | ···················· | ···················· | ···················· | |
| V | 6784 hCoV-19/Japan/PG-7344/2020\|EPI_ISL_688447\|2020-08 | ···················· | ···················· | ···················· | ···················· | ···················· | ···················· | |
| V | 6785 hCoV-19/Japan/PG-7345/2020\|EPI_ISL_688448\|2020-08 | ···················· | ···················· | ···················· | ···················· | ···················· | ···················· | |
| V | 6786 hCoV-19/Japan/PG-7346/2020\|EPI_ISL_688449\|2020-08 | ···················· | ···················· | ···················· | ···················· | ···················· | ···················· | |
| V | 6787 hCoV-19/Japan/PG-7348/2020\|EPI_ISL_688450\|2020-08 | ···················· | ···················· | ···················· | ···················· | ···················· | ···················· | |
| V | 6788 hCoV-19/Japan/PG-7349/2020\|EPI_ISL_688451\|2020-08 | ···················· | ···················· | ···················· | ···················· | ···················· | ···················· | |
| V | 6789 hCoV-19/Japan/PG-7350/2020\|EPI_ISL_688452\|2020-08 | ···················· | ···················· | ···················· | ···················· | ···················· | ···················· | |
| V | 6790 hCoV-19/Japan/PG-7351/2020\|EPI_ISL_688453\|2020-08 | ···················· | ···················· | ···················· | ···················· | ···················· | ···················· | |
| V | 6791 hCoV-19/Japan/PG-7352/2020\|EPI_ISL_688454\|2020-08 | ···················· | ···················· | ···················· | ···················· | ···················· | ···················· | |
| V | 6792 hCoV-19/Japan/PG-7353/2020\|EPI_ISL_688455\|2020-08 | ···················· | ···················· | ···················· | ···················· | ···················· | ···················· | |
| V | 6793 hCoV-19/Japan/PG-7354/2020\|EPI_ISL_688456\|2020-08 | ···················· | ···················· | ···················· | ···················· | ···················· | ···················· | |
| V | 6794 hCoV-19/Japan/PG-7355/2020\|EPI_ISL_688457\|2020-08 | ···················· | ···················· | ···················· | ···················· | ···················· | ···················· | |
| V | 6795 hCoV-19/Japan/PG-7356/2020\|EPI_ISL_688458\|2020-08 | ···················· | ···················· | ···················· | ···················· | ···················· | ···················· | |
| V | 6796 hCoV-19/Japan/PG-7357/2020\|EPI_ISL_688459\|2020-08 | ···················· | ···················· | ···················· | ···················· | ···················· | NNN················· | 1 |
| V | 6797 hCoV-19/Japan/PG-7358/2020\|EPI_ISL_688460\|2020-08 | ···················· | ···················· | ···················· | ···················· | ···················· | ···················· | |
| V | 6798 hCoV-19/Japan/PG-7359/2020\|EPI_ISL_688461\|2020-08 | ···················· | ···················· | ···················· | ···················· | ···················· | ···················· | |
| V | 6799 hCoV-19/Japan/PG-7360/2020\|EPI_ISL_688462\|2020-08 | ···················· | ···················· | ···················· | ···················· | ···················· | ···················· | |
| V | 6800 hCoV-19/Japan/PG-7361/2020\|EPI_ISL_688463\|2020-08 | ···················· | ···················· | ···················· | ···················· | ···················· | ···················· | |
| V | 6801 hCoV-19/Japan/PG-7362/2020\|EPI_ISL_688464\|2020-08 | ···················· | ···················· | ···················· | ···················· | ···················· | ···················· | |
| V | 6802 hCoV-19/Japan/PG-7363/2020\|EPI_ISL_688465\|2020-08 | ···················· | ···················· | ···················· | ···················· | ···················· | ···················· | |
| V | 6803 hCoV-19/Japan/PG-7364/2020\|EPI_ISL_688466\|2020-08 | ···················· | ···················· | ···················· | ···················· | ···················· | ···················· | |
| V | 6804 hCoV-19/Japan/PG-7365/2020\|EPI_ISL_688467\|2020-08 | ···················· | ···················· | ···················· | ···················· | ···················· | ···················· | |
| V | 6805 hCoV-19/Japan/PG-7366/2020\|EPI_ISL_688468\|2020-06 | ···················· | ···················· | ···················· | ···················· | ···················· | ···················· | |
| V | 6806 hCoV-19/Japan/PG-7367/2020\|EPI_ISL_688469\|2020-06 | ···················· | ···················· | ···················· | ···················· | ···················· | ···················· | |
| V | 6807 hCoV-19/Japan/PG-7368/2020\|EPI_ISL_688470\|2020-06 | ···················· | ···················· | ···················· | ···················· | ···················· | ···················· | |
| V | 6808 hCoV-19/Japan/PG-7369/2020\|EPI_ISL_688471\|2020-06 | ···················· | ···················· | ···················· | ···················· | ···················· | ···················· | |
| V | 6809 hCoV-19/Japan/PG-7370/2020\|EPI_ISL_688472\|2020-06 | ···················· | ···················· | ···················· | ···················· | ···················· | ···················· | |
| V | 6810 hCoV-19/Japan/PG-7371/2020\|EPI_ISL_688473\|2020-06 | ···················· | ···················· | ···················· | ···················· | ···················· | ···················· | |
| V | 6811 hCoV-19/Japan/PG-7372/2020\|EPI_ISL_688474\|2020-06 | ···················· | ···················· | ···················· | ···················· | ···················· | ···················· | |
| V | 6812 hCoV-19/Japan/PG-7373/2020\|EPI_ISL_688475\|2020-06 | ···················· | ···················· | ···················· | ···················· | ···················· | ···················· | |
| V | 6813 hCoV-19/Japan/PG-7374/2020\|EPI_ISL_688476\|2020-06 | ···················· | ···················· | ···················· | ···················· | ···················· | ···················· | |
| V | 6814 hCoV-19/Japan/PG-7375/2020\|EPI_ISL_688477\|2020-06 | ···················· | ···················· | ···················· | ···················· | ···················· | ···················· | |
| V | 6815 hCoV-19/Japan/PG-7376/2020\|EPI_ISL_688478\|2020-06 | ···················· | ···················· | ···················· | ···················· | ···················· | ···················· | |
| V | 6816 hCoV-19/Japan/PG-7377/2020\|EPI_ISL_688479\|2020-07 | ···················· | ···················· | ···················· | ···················· | ···················· | ···················· | |
| V | 6817 hCoV-19/Japan/PG-7398/2020\|EPI_ISL_688500\|2020-07 | ···················· | ···················· | ···················· | ···················· | ···················· | ···················· | |
| V | 6818 hCoV-19/Japan/PG-7399/2020\|EPI_ISL_688501\|2020-07 | ···················· | ···················· | ···················· | ···················· | ···················· | ···················· | |
| V | 6819 hCoV-19/Japan/PG-7400/2020\|EPI_ISL_688502\|2020-07 | ···················· | ···················· | ···················· | ···················· | ···················· | ···················· | |
| V | 6820 hCoV-19/Japan/PG-7401/2020\|EPI_ISL_688503\|2020-07 | ···················· | ···················· | ···················· | ···················· | ···················· | ···················· | |
| V | 6821 hCoV-19/Japan/PG-7402/2020\|EPI_ISL_688504\|2020-07 | ···················· | ···················· | ···················· | ···················· | ···················· | ···················· | |
| V | 6822 hCoV-19/Japan/PG-7403/2020\|EPI_ISL_688505\|2020-07 | ···················· | ···················· | ···················· | ···················· | ···················· | ···················· | |
| V | 6823 hCoV-19/Japan/PG-7404/2020\|EPI_ISL_688506\|2020-07 | ···················· | ···················· | ···················· | ···················· | ···················· | ···················· | |
| V | 6824 hCoV-19/Japan/PG-7405/2020\|EPI_ISL_688507\|2020-07 | ···················· | ···················· | ···················· | ···················· | ···················· | ···················· | |
| V | 6825 hCoV-19/Japan/PG-7406/2020\|EPI_ISL_688508\|2020-07 | ···················· | ···················· | ···················· | ···················· | ···················· | ···················· | |
| V | 6826 hCoV-19/Japan/PG-7407/2020\|EPI_ISL_688509\|2020-07 | ···················· | ···················· | ···················· | ···················· | ···················· | ···················· | |
| V | 6827 hCoV-19/Japan/PG-7408/2020\|EPI_ISL_688510\|2020-07 | ···················· | ···················· | ···················· | ···················· | ···················· | ···················· | |
| V | 6828 hCoV-19/Japan/PG-7409/2020\|EPI_ISL_688511\|2020-07 | ···················· | ···················· | ···················· | ···················· | ···················· | ···················· | |
| V | 6829 hCoV-19/Japan/PG-7410/2020\|EPI_ISL_688512\|2020-07 | ···················· | ···················· | ···················· | ···················· | ···················· | ···················· | |
| V | 6830 hCoV-19/Japan/PG-7411/2020\|EPI_ISL_688513\|2020-07 | ···················· | ···················· | ···················· | ···················· | ···················· | ···················· | |
| V | 6831 hCoV-19/Japan/PG-4587/2020\|EPI_ISL_689724\|2020-08 | ···················· | ···················· | ···················· | ···················· | ···················· | ···················· | |
| V | 6832 hCoV-19/Japan/PG-4588/2020\|EPI_ISL_689725\|2020-08 | ···················· | ···················· | ···················· | ···················· | ···················· | ···················· | |
| V | 6833 hCoV-19/Japan/PG-4589/2020\|EPI_ISL_689726\|2020-08 | ···················· | ···················· | ···················· | ···················· | ···················· | ···················· | |
| V | 6834 hCoV-19/Japan/PG-4590/2020\|EPI_ISL_689727\|2020-08 | ···················· | ···················· | ···················· | ···················· | ···················· | ···················· | |
| V | 6835 hCoV-19/Japan/PG-4591/2020\|EPI_ISL_689728\|2020-08 | ···················· | ···················· | ···················· | ···················· | ···················· | ···················· | |
| V | 6836 hCoV-19/Japan/PG-4592/2020\|EPI_ISL_689729\|2020-08 | ···················· | ···················· | ···················· | ···················· | ···················· | ···················· | |
| V | 6837 hCoV-19/Japan/PG-4593/2020\|EPI_ISL_689730\|2020-08 | ···················· | ···················· | ···················· | ···················· | ···················· | ···················· | |
| V | 6838 hCoV-19/Japan/PG-4594/2020\|EPI_ISL_689731\|2020-08 | ···················· | ···················· | ···················· | ···················· | ···················· | ···················· | |
| V | 6839 hCoV-19/Japan/PG-4595/2020\|EPI_ISL_689732\|2020-08 | ···················· | ···················· | ···················· | ···················· | ···················· | ···················· | |
| V | 6840 hCoV-19/Japan/PG-4596/2020\|EPI_ISL_689733\|2020-08 | ···················· | ···················· | ···················· | ···················· | ···················· | ···················· | |
| V | 6841 hCoV-19/Japan/PG-4597/2020\|EPI_ISL_689734\|2020-08 | ···················· | ···················· | ···················· | ···················· | ···················· | ···················· | |
| V | 6842 hCoV-19/Japan/PG-4598/2020\|EPI_ISL_689735\|2020-08 | ···················· | ···················· | ···················· | ···················· | ···················· | ···················· | |
| V | 6843 hCoV-19/Japan/PG-4599/2020\|EPI_ISL_689736\|2020-08 | ···················· | ···················· | ···················· | ···················· | ···················· | ···················· | |
| V | 6844 hCoV-19/Japan/PG-4600/2020\|EPI_ISL_689737\|2020-08 | ···················· | ···················· | ···················· | ···················· | ···················· | ···················· | |
| V | 6845 hCoV-19/Japan/PG-4601/2020\|EPI_ISL_689738\|2020-08 | ···················· | ···················· | ···················· | ···················· | ···················· | ···················· | |
| V | 6846 hCoV-19/Japan/PG-4602/2020\|EPI_ISL_689739\|2020-08 | ···················· | ···················· | ···················· | ···················· | ···················· | ···················· | |
| V | 6847 hCoV-19/Japan/PG-4603/2020\|EPI_ISL_689740\|2020-08 | ···················· | ···················· | ···················· | ···················· | ···················· | ···················· | |
| V | 6848 hCoV-19/Japan/PG-4604/2020\|EPI_ISL_689741\|2020-08 | ···················· | ···················· | ···················· | ···················· | ···················· | ···················· | |
| V | 6849 hCoV-19/Japan/PG-4605/2020\|EPI_ISL_689742\|2020-08 | ···················· | ···················· | ···················· | ···················· | ···················· | ···················· | |
| V | 6850 hCoV-19/Japan/PG-4606/2020\|EPI_ISL_689743\|2020-08 | ···················· | ···················· | ···················· | ···················· | ···················· | ···················· | |
| V | 6851 hCoV-19/Japan/PG-4607/2020\|EPI_ISL_689744\|2020-08 | ···················· | ···················· | ···················· | ···················· | ···················· | ···················· | |
| V | 6852 hCoV-19/Japan/PG-4608/2020\|EPI_ISL_689745\|2020-08 | ···················· | ···················· | ···················· | ···················· | ···················· | ···················· | |

| | | | | | | | | |
|---|---|---|---|---|---|---|---|---|
| V | 6853 hCoV-19/Japan/PG-4609/2020\|EPI_ISL_689746\|2020-08 | .................... | .................... | ..................... | .................... | .................... | .................... | |
| V | 6854 hCoV-19/Japan/PG-4610/2020\|EPI_ISL_689747\|2020-08 | .................... | .................... | ..................... | .................... | .................... | .................... | |
| V | 6855 hCoV-19/Japan/PG-4611/2020\|EPI_ISL_689748\|2020-08 | .................... | .................... | ..................... | .................... | .................... | .................... | |
| V | 6856 hCoV-19/Japan/PG-4612/2020\|EPI_ISL_689749\|2020-08 | .................... | .................... | ..................... | .................... | .................... | .................... | |
| V | 6857 hCoV-19/Japan/PG-4613/2020\|EPI_ISL_689750\|2020-08 | .................... | .................... | ..................... | .................... | .................... | .................... | |
| V | 6858 hCoV-19/Japan/PG-4704/2020\|EPI_ISL_689841\|2020-08 | .................... | .................... | ..................... | .................... | .................... | .................... | |
| V | 6859 hCoV-19/Japan/PG-4705/2020\|EPI_ISL_689842\|2020-07 | .................... | .................... | ..................... | .................... | .................... | .................... | |
| V | 6860 hCoV-19/Japan/PG-4706/2020\|EPI_ISL_689843\|2020-07 | .................... | .................... | ..................... | .................... | .................... | .................... | |
| V | 6861 hCoV-19/Japan/PG-4707/2020\|EPI_ISL_689844\|2020-07 | .................... | .................... | ..................... | .................... | .................... | .................... | |
| V | 6862 hCoV-19/Japan/PG-2576/2020\|EPI_ISL_690710\|2020-03 | .................... | .................... | ..................... | .................... | .................... | .................... | |
| V | 6863 hCoV-19/Japan/PG-2577/2020\|EPI_ISL_690711\|2020-04 | .................... | .................... | ..................... | .................... | .................... | .................... | |
| V | 6864 hCoV-19/Japan/PG-2578/2020\|EPI_ISL_690712\|2020-04 | .................... | .................... | ..................... | .................... | .................... | .................... | |
| V | 6865 hCoV-19/Japan/PG-2579/2020\|EPI_ISL_690713\|2020-04 | .................... | .................... | ..................... | .................... | .................... | .................... | |
| V | 6866 hCoV-19/Japan/PG-2580/2020\|EPI_ISL_690714\|2020-04 | .................... | .................... | ..................... | .................... | .................... | .................... | |
| V | 6867 hCoV-19/Japan/PG-2581/2020\|EPI_ISL_690715\|2020-04 | .................... | .................... | ..................... | .................... | .................... | .................... | |
| V | 6868 hCoV-19/Japan/PG-2582/2020\|EPI_ISL_690716\|2020-04 | .................... | .................... | ..................... | .................... | .................... | .................... | |
| V | 6869 hCoV-19/Japan/PG-2583/2020\|EPI_ISL_690717\|2020-04 | .................... | .................... | ..................... | .................... | .................... | .................... | |
| V | 6870 hCoV-19/Japan/PG-2584/2020\|EPI_ISL_690718\|2020-04 | .................... | .................... | ..................... | .................... | .................... | .................... | |
| V | 6871 hCoV-19/Japan/PG-2585/2020\|EPI_ISL_690719\|2020-04 | .................... | .................... | ..................... | .................... | .................... | .................... | |
| V | 6872 hCoV-19/Japan/PG-2586/2020\|EPI_ISL_690720\|2020-04 | .................... | .................... | ..................... | .................... | .................... | .................... | |
| V | 6873 hCoV-19/Japan/PG-2587/2020\|EPI_ISL_690721\|2020-04 | .................... | .................... | ..................... | .................... | .................... | .................... | |
| V | 6874 hCoV-19/Japan/PG-2588/2020\|EPI_ISL_690722\|2020-04 | .................... | .................... | ..................... | .................... | .................... | .................... | |
| V | 6875 hCoV-19/Japan/PG-2589/2020\|EPI_ISL_690723\|2020-04 | .................... | .................... | ..................... | .................... | .................... | .................... | |
| V | 6876 hCoV-19/Japan/PG-2590/2020\|EPI_ISL_690724\|2020-04 | .................... | .................... | ..................... | .................... | .................... | .................... | |
| V | 6877 hCoV-19/Japan/PG-2591/2020\|EPI_ISL_690725\|2020-04 | .................... | .................... | ..................... | .................... | .................... | .................... | |
| V | 6878 hCoV-19/Japan/PG-2592/2020\|EPI_ISL_690726\|2020-04 | .................... | .................... | ..................... | .................... | .................... | .................... | |
| V | 6879 hCoV-19/Japan/PG-2593/2020\|EPI_ISL_690727\|2020-04 | .................... | .................... | ..................... | .................... | .................... | .................... | |
| V | 6880 hCoV-19/Japan/PG-2594/2020\|EPI_ISL_690728\|2020-04 | .................... | .................... | ..................... | .................... | .................... | .................... | |
| V | 6881 hCoV-19/Japan/PG-2595/2020\|EPI_ISL_690729\|2020-04 | .................... | .................... | ..................... | .................... | .................... | .................... | |
| V | 6882 hCoV-19/Japan/PG-2596/2020\|EPI_ISL_690730\|2020-04 | .................... | .................... | ..................... | .................... | .................... | .................... | |
| V | 6883 hCoV-19/Japan/PG-2597/2020\|EPI_ISL_690731\|2020-04 | .................... | .................... | ..................... | .................... | .................... | .................... | |
| V | 6884 hCoV-19/Japan/PG-2598/2020\|EPI_ISL_690732\|2020-04 | .................... | .................... | ..................... | .................... | .................... | .................... | |
| V | 6885 hCoV-19/Japan/PG-2599/2020\|EPI_ISL_690733\|2020-04 | .................... | .................... | ..................... | .................... | .................... | .................... | |
| V | 6886 hCoV-19/Japan/PG-2600/2020\|EPI_ISL_690734\|2020-04 | .................... | .................... | ..................... | .................... | .................... | .................... | |
| V | 6887 hCoV-19/Japan/PG-2601/2020\|EPI_ISL_690735\|2020-04 | .................... | .................... | ..................... | .................... | .................... | .................... | |
| V | 6888 hCoV-19/Japan/PG-2602/2020\|EPI_ISL_690736\|2020-04 | .................... | .................... | ..................... | .................... | .................... | .................... | |
| V | 6889 hCoV-19/Japan/PG-2603/2020\|EPI_ISL_690737\|2020-04 | .................... | .................... | ..................... | .................... | .................... | .................... | |
| V | 6890 hCoV-19/Japan/PG-2604/2020\|EPI_ISL_690738\|2020-04 | .................... | .................... | ..................... | .................... | .................... | .................... | |
| V | 6891 hCoV-19/Japan/PG-2605/2020\|EPI_ISL_690739\|2020-04 | .................... | .................... | ..................... | .................... | .................... | .................... | |
| V | 6892 hCoV-19/Japan/PG-2659/2020\|EPI_ISL_690791\|2020-04 | .................... | .................... | ..................... | .................... | .................... | .................... | |
| V | 6893 hCoV-19/Japan/PG-2660/2020\|EPI_ISL_690792\|2020-04 | .................... | .................... | ..................... | .................... | .................... | .................... | |
| V | 6894 hCoV-19/Japan/PG-2661/2020\|EPI_ISL_690793\|2020-04 | .................... | .................... | ..................... | .................... | .................... | .................... | |
| V | 6895 hCoV-19/Japan/PG-2662/2020\|EPI_ISL_690794\|2020-06 | .................... | .................... | ..................... | .................... | .................... | .................... | |
| V | 6896 hCoV-19/Japan/PG-2663/2020\|EPI_ISL_690795\|2020-06 | .................... | .................... | ..................... | .................... | .................... | .................... | |
| V | 6897 hCoV-19/Japan/PG-2664/2020\|EPI_ISL_690796\|2020-06 | .................... | .................... | ..................... | .................... | .................... | .................... | |
| V | 6898 hCoV-19/Japan/PG-2665/2020\|EPI_ISL_690797\|2020-06 | .................... | .................... | ..................... | .................... | .................... | .................... | |
| V | 6899 hCoV-19/Japan/PG-2666/2020\|EPI_ISL_690798\|2020-06 | .................... | .................... | ..................... | .................... | .................... | .................... | |
| V | 6900 hCoV-19/Japan/PG-2667/2020\|EPI_ISL_690799\|2020-06 | .................... | .................... | ..................... | .................... | .................... | .................... | |
| V | 6901 hCoV-19/USA/MA-MASPHL-00625/2020\|EPI_ISL_693000\|20 | .................... | .................... | ..................... | .................... | .................... | .................... | |
| V | 6902 hCoV-19/USA/MA-MASPHL-00622/2020\|EPI_ISL_693003\|20 | .................... | ...................A | ..................... | .................... | .................... | .................... | 1 |
| V | 6903 hCoV-19/USA/MA-MASPHL-00621/2020\|EPI_ISL_693004\|20 | .................... | .................... | ...........T......... | .................... | .................... | .................... | |
| V | 6904 hCoV-19/USA/MA-MASPHL-00615/2020\|EPI_ISL_693006\|20 | .................... | .................... | ..................... | .................... | .................... | .................... | |
| V | 6905 hCoV-19/USA/MA-MASPHL-00612/2020\|EPI_ISL_693008\|20 | .................... | .................... | ..................... | .................... | .................... | .................... | |
| V | 6906 hCoV-19/USA/MA-MASPHL-00611/2020\|EPI_ISL_693009\|20 | .................... | .................... | ..................... | .................... | .................... | .................... | |
| V | 6907 hCoV-19/USA/MA-MASPHL-00606/2020\|EPI_ISL_693013\|20 | .................... | .................... | ..................... | .................... | .................... | .................... | |
| V | 6908 hCoV-19/USA/MA-MASPHL-00593/2020\|EPI_ISL_693016\|20 | .................... | .................... | ..................... | .................... | .................... | .................... | |
| V | 6909 hCoV-19/USA/MA-MASPHL-00591/2020\|EPI_ISL_693018\|20 | .................... | .................... | ..................... | .................... | .................... | .................... | |
| V | 6910 hCoV-19/USA/MA-MASPHL-00588/2020\|EPI_ISL_693019\|20 | .................... | .................... | ..................... | .................... | .................... | .................... | |
| V | 6911 hCoV-19/USA/MA-MASPHL-00583/2020\|EPI_ISL_693021\|20 | .................... | .................... | ..................... | .................... | .................... | .................... | |
| V | 6912 hCoV-19/USA/MA-MASPHL-00578/2020\|EPI_ISL_693023\|20 | .................... | .................... | ..................... | .................... | .................... | .................... | |
| V | 6913 hCoV-19/Japan/PG-1292/2020\|EPI_ISL_684900\|2020-03 | .................... | .................... | ..................... | .................... | .................... | .................... | |
| V | 6914 hCoV-19/Japan/PG-1293/2020\|EPI_ISL_684901\|2020-03 | .................... | .................... | ..................... | .................... | .................... | .................... | |
| V | 6915 hCoV-19/Japan/PG-1294/2020\|EPI_ISL_684902\|2020-03 | .................... | .................... | ..................... | .................... | .................... | .................... | |
| V | 6916 hCoV-19/Japan/PG-1295/2020\|EPI_ISL_684903\|2020-03 | .................... | .................... | ..................... | .................... | .................... | .................... | |
| V | 6917 hCoV-19/Japan/PG-1296/2020\|EPI_ISL_684904\|2020-03 | .................... | .................... | ..................... | .................... | .................... | .................... | |
| V | 6918 hCoV-19/Japan/PG-1297/2020\|EPI_ISL_684905\|2020-03 | .................... | .................... | ..................... | .................... | .................... | .................... | |
| V | 6919 hCoV-19/Japan/PG-1298/2020\|EPI_ISL_684906\|2020-03 | .................... | .................... | ..................... | .................... | .................... | .................... | |
| V | 6920 hCoV-19/Japan/PG-1299/2020\|EPI_ISL_684907\|2020-03 | .................... | .................... | ..................... | .................... | .................... | .................... | |
| V | 6921 hCoV-19/Japan/PG-1300/2020\|EPI_ISL_684908\|2020-03 | .................... | .................... | ..................... | .................... | .................... | .................... | |
| V | 6922 hCoV-19/Japan/PG-1301/2020\|EPI_ISL_684909\|2020-03 | .................... | .................... | ..................... | .................... | .................... | .................... | |
| V | 6923 hCoV-19/Japan/PG-1302/2020\|EPI_ISL_684910\|2020-04 | .................... | .................... | ..................... | .................... | .................... | .................... | |
| V | 6924 hCoV-19/Japan/PG-1303/2020\|EPI_ISL_684911\|2020-04 | .................... | .................... | ..................... | .................... | .................... | .................... | |
| V | 6925 hCoV-19/Japan/PG-1304/2020\|EPI_ISL_684912\|2020-04 | .................... | .................... | ..................... | .................... | .................... | .................... | |
| V | 6926 hCoV-19/Japan/PG-1305/2020\|EPI_ISL_684913\|2020-04 | .................... | .................... | ..................... | .................... | .................... | .................... | |
| V | 6927 hCoV-19/Japan/PG-1306/2020\|EPI_ISL_684914\|2020-04 | .................... | .................... | ..................... | .................... | .................... | .................... | |
| V | 6928 hCoV-19/Japan/PG-1307/2020\|EPI_ISL_684915\|2020-04 | .................... | .................... | ..................... | .................... | .................... | .................... | |
| V | 6929 hCoV-19/Japan/PG-1308/2020\|EPI_ISL_684916\|2020-04 | .................... | .................... | ..................... | .................... | .................... | .................... | |
| V | 6930 hCoV-19/Japan/PG-1309/2020\|EPI_ISL_684917\|2020-04 | .................... | .................... | ..................... | .................... | .................... | .................... | |
| V | 6931 hCoV-19/Japan/PG-1310/2020\|EPI_ISL_684918\|2020-04 | .................... | .................... | ..................... | .................... | .................... | .................... | |
| V | 6932 hCoV-19/Japan/PG-1311/2020\|EPI_ISL_684919\|2020-04 | .................... | .................... | ..................... | .................... | .................... | .................... | |

| | | | | | | | |
|---|---|---|---|---|---|---|---|
| V | 6933 hCoV-19/Japan/PG-1312/2020\|EPI_ISL_684920\|2020-04 | .................... | .................... | ..................... | .................... | .................... | .................... |
| V | 6934 hCoV-19/Japan/PG-1313/2020\|EPI_ISL_684921\|2020-04 | .................... | .................... | ..................... | .................... | .................... | .................... |
| V | 6935 hCoV-19/Japan/PG-1315/2020\|EPI_ISL_684923\|2020-04 | .................... | .................... | ..................... | .................... | .................... | .................... |
| V | 6936 hCoV-19/Japan/PG-1316/2020\|EPI_ISL_684924\|2020-04 | .................... | .................... | ..................... | .................... | .................... | .................... |
| V | 6937 hCoV-19/Japan/PG-1754/2020\|EPI_ISL_685360\|2020-04 | .................... | .................... | ..................... | .................... | .................... | .................... |
| V | 6938 hCoV-19/Japan/PG-1755/2020\|EPI_ISL_685361\|2020-04 | .................... | .................... | ..................... | .................... | .................... | .................... |
| V | 6939 hCoV-19/Japan/PG-1756/2020\|EPI_ISL_685362\|2020-04 | .................... | .................... | ..................... | .................... | .................... | .................... |
| V | 6940 hCoV-19/Japan/PG-1757/2020\|EPI_ISL_685363\|2020-04 | .................... | .................... | ..................... | .................... | .................... | .................... |
| V | 6941 hCoV-19/Japan/PG-1758/2020\|EPI_ISL_685364\|2020-04 | .................... | .................... | ..................... | .................... | .................... | .................... |
| V | 6942 hCoV-19/Japan/PG-1759/2020\|EPI_ISL_685365\|2020-04 | .................... | .................... | ..................... | .................... | .................... | .................... |
| V | 6943 hCoV-19/Japan/PG-1760/2020\|EPI_ISL_685366\|2020-04 | .................... | .................... | ..................... | .................... | .................... | .................... |
| V | 6944 hCoV-19/Japan/PG-1761/2020\|EPI_ISL_685367\|2020-04 | .................... | .................... | ..................... | .................... | .................... | .................... |
| V | 6945 hCoV-19/England/204691665/2020\|EPI_ISL_686600\|2020 | .................... | .................... | ..................... | .................... | .................... | .................... |
| V | 6946 hCoV-19/England/204720047/2020\|EPI_ISL_686601\|2020 | .................... | .................... | ..................... | .................... | .................... | .................... |
| V | 6947 hCoV-19/England/204720209/2020\|EPI_ISL_686602\|2020 | .................... | .................... | ..................... | .................... | .................... | .................... |
| V | 6948 hCoV-19/England/204720235/2020\|EPI_ISL_686603\|2020 | .................... | .................... | ..................... | .................... | .................... | .................... |
| V | 6949 hCoV-19/England/204720512/2020\|EPI_ISL_686604\|2020 | .................... | .................... | ..................... | .................... | .................... | .................... |
| V | 6950 hCoV-19/England/204720848/2020\|EPI_ISL_686605\|2020 | .................... | .A.......A.......... | ..................... | .................... | .................... | .................... |
| V | 6951 hCoV-19/England/204720858/2020\|EPI_ISL_686606\|2020 | .................... | .................... | ..................... | .................... | .................... | .................... |
| V | 6952 hCoV-19/England/204720866/2020\|EPI_ISL_686607\|2020 | .................... | .................... | ..................... | .................... | .................... | .................... |
| V | 6953 hCoV-19/England/204720888/2020\|EPI_ISL_686608\|2020 | .................... | .................... | ..................... | .................... | .................... | .................... |
| V | 6954 hCoV-19/England/204720936/2020\|EPI_ISL_686609\|2020 | .................... | .................... | ..................... | .................... | .................... | .................... |
| V | 6955 hCoV-19/Japan/PG-7589/2020\|EPI_ISL_686691\|2020-07 | .................... | .................... | ..................... | .................... | .................... | .................... |
| V | 6956 hCoV-19/Japan/PG-7590/2020\|EPI_ISL_686692\|2020-07 | .................... | .................... | ..................... | .................... | .................... | .................... |
| V | 6957 hCoV-19/Japan/PG-7591/2020\|EPI_ISL_686693\|2020-07 | .................... | .................... | ..................... | .................... | .................... | .................... |
| V | 6958 hCoV-19/Japan/PG-7592/2020\|EPI_ISL_686694\|2020-07 | .................... | .................... | ..................... | .................... | .................... | .................... |
| V | 6959 hCoV-19/Japan/PG-7593/2020\|EPI_ISL_686695\|2020-07 | ........T........... | .................... | ..................... | .................... | .................... | .................... |
| V | 6960 hCoV-19/Japan/PG-7594/2020\|EPI_ISL_686696\|2020-07 | .................... | .................... | ..................... | .................... | .................... | .................... |
| V | 6961 hCoV-19/Japan/PG-7595/2020\|EPI_ISL_686697\|2020-07 | .................... | .................... | ..................... | .................... | .................... | .................... |
| V | 6962 hCoV-19/Japan/PG-7596/2020\|EPI_ISL_686698\|2020-07 | .................... | .................... | ..................... | .................... | .................... | .................... |
| V | 6963 hCoV-19/Japan/PG-8049/2020\|EPI_ISL_687150\|2020-09 | .................... | .................... | ..................... | .................... | .................... | .................... |
| V | 6964 hCoV-19/Japan/PG-8050/2020\|EPI_ISL_687151\|2020-09 | .................... | .................... | ..................... | .................... | .................... | .................... |
| V | 6965 hCoV-19/Japan/PG-8051/2020\|EPI_ISL_687152\|2020-08 | .................... | .................... | ..................... | .................... | .................... | .................... |
| V | 6966 hCoV-19/Japan/PG-8052/2020\|EPI_ISL_687153\|2020-08 | .................... | .................... | ..................... | .................... | .................... | .................... |
| V | 6967 hCoV-19/Japan/PG-8053/2020\|EPI_ISL_687154\|2020-08 | .................... | .................... | ..................... | .................... | .................... | .................... |
| V | 6968 hCoV-19/Japan/PG-8054/2020\|EPI_ISL_687155\|2020-08 | .................... | .................... | ..................... | .................... | .................... | .................... |
| V | 6969 hCoV-19/Japan/PG-8055/2020\|EPI_ISL_687156\|2020-08 | .................... | .................... | ..................... | .................... | .................... | .................... |
| V | 6970 hCoV-19/Japan/PG-8056/2020\|EPI_ISL_687157\|2020-08 | .................... | .................... | ..................... | .................... | .................... | .................... |
| V | 6971 hCoV-19/Japan/PG-8057/2020\|EPI_ISL_687158\|2020-09 | .................... | .................... | ..................... | .................... | .................... | .................... |
| V | 6972 hCoV-19/Japan/PG-8058/2020\|EPI_ISL_687159\|2020-09 | .................... | .................... | ..................... | .................... | .................... | .................... |
| V | 6973 hCoV-19/Japan/PG-8059/2020\|EPI_ISL_687160\|2020-09 | .................... | .................... | ..................... | .................... | .................... | .................... |
| V | 6974 hCoV-19/Japan/PG-8060/2020\|EPI_ISL_687161\|2020-09 | .................... | .................... | ..................... | .................... | .................... | .................... |
| V | 6975 hCoV-19/Japan/PG-8061/2020\|EPI_ISL_687162\|2020-09 | .................... | .................... | ..................... | .................... | .................... | .................... |
| V | 6976 hCoV-19/Japan/PG-8062/2020\|EPI_ISL_687163\|2020-07 | .................... | .................... | ..................... | .................... | .................... | .................... |
| V | 6977 hCoV-19/Japan/PG-8063/2020\|EPI_ISL_687164\|2020-07 | .................... | .................... | ..................... | .................... | .................... | .................... |
| V | 6978 hCoV-19/Japan/PG-8064/2020\|EPI_ISL_687165\|2020-07 | .................... | .................... | ..................... | .................... | .................... | .................... |
| V | 6979 hCoV-19/Japan/PG-8065/2020\|EPI_ISL_687166\|2020-07 | .................... | .................... | ..................... | .................... | .................... | .................... |
| V | 6980 hCoV-19/Japan/PG-8071/2020\|EPI_ISL_687172\|2020-07 | .................... | .................... | ..................... | .................... | .................... | .................... |
| V | 6981 hCoV-19/Japan/PG-8072/2020\|EPI_ISL_687173\|2020-07 | .................... | .................... | ..................... | .................... | .................... | .................... |
| V | 6982 hCoV-19/Japan/PG-8073/2020\|EPI_ISL_687174\|2020-07 | .................... | .................... | ..................... | .................... | .................... | .................... |
| V | 6983 hCoV-19/Japan/PG-8076/2020\|EPI_ISL_687177\|2020-08 | .................... | .................... | ..................... | .................... | .................... | .................... |
| V | 6984 hCoV-19/Japan/PG-8078/2020\|EPI_ISL_687179\|2020-08 | .................... | .................... | ..................... | .................... | .................... | .................... |
| V | 6985 hCoV-19/Japan/PG-8080/2020\|EPI_ISL_687181\|2020-08 | .................... | .................... | ..................... | .................... | .................... | .................... |
| V | 6986 hCoV-19/Japan/PG-8081/2020\|EPI_ISL_687182\|2020-08 | .................... | .................... | ..................... | .................... | .................... | .................... |
| V | 6987 hCoV-19/Japan/PG-8083/2020\|EPI_ISL_687184\|2020-08 | .................... | .................... | ..................... | .................... | .................... | .................... |
| V | 6988 hCoV-19/Japan/PG-8084/2020\|EPI_ISL_687185\|2020-08 | .................... | .................... | ..................... | .................... | .................... | .................... |
| V | 6989 hCoV-19/Japan/PG-8088/2020\|EPI_ISL_687188\|2020-08 | .................... | .................... | ..................... | .................... | .................... | .................... |
| V | 6990 hCoV-19/Japan/PG-8089/2020\|EPI_ISL_687189\|2020-08 | .................... | .................... | ..................... | .................... | .................... | .................... |
| V | 6991 hCoV-19/Japan/PG-8090/2020\|EPI_ISL_687190\|2020-08 | .................... | .................... | ..................... | .................... | .................... | .................... |
| V | 6992 hCoV-19/Japan/PG-8092/2020\|EPI_ISL_687191\|2020-08 | .................... | .................... | ..................... | .................... | .................... | .................... |
| V | 6993 hCoV-19/Japan/PG-8093/2020\|EPI_ISL_687192\|2020-08 | .................... | .................... | ..................... | .................... | .................... | .................... |
| V | 6994 hCoV-19/Japan/PG-8094/2020\|EPI_ISL_687193\|2020-08 | .................... | .................... | ..................... | .................... | .................... | .................... |
| V | 6995 hCoV-19/Japan/PG-8095/2020\|EPI_ISL_687194\|2020-08 | .................... | .................... | ..................... | .................... | .................... | .................... |
| V | 6996 hCoV-19/Japan/PG-8096/2020\|EPI_ISL_687195\|2020-08 | .................... | .................... | ..................... | .................... | .................... | .................... |
| V | 6997 hCoV-19/Japan/PG-8097/2020\|EPI_ISL_687196\|2020-08 | .................... | .................... | ..................... | .................... | .................... | .................... |
| V | 6998 hCoV-19/Japan/PG-8098/2020\|EPI_ISL_687197\|2020-08 | .................... | .................... | ..................... | .................... | .................... | .................... |
| V | 6999 hCoV-19/Japan/PG-8099/2020\|EPI_ISL_687198\|2020-08 | .................... | .................... | ..................... | .................... | .................... | .................... |
| V | 7000 hCoV-19/Japan/PG-8100/2020\|EPI_ISL_687199\|2020-08 | .................... | .................... | ..................... | .................... | .................... | .................... |
| V | 7001 hCoV-19/Japan/PG-6791/2020\|EPI_ISL_687900\|2020-08 | .................... | .................... | ..................... | .................... | .................... | .................... |
| V | 7002 hCoV-19/Japan/PG-6792/2020\|EPI_ISL_687901\|2020-08 | .................... | .................... | ..................... | .................... | .................... | .................... |
| V | 7003 hCoV-19/Japan/PG-6793/2020\|EPI_ISL_687902\|2020-08 | .................... | .................... | ..................... | .................... | .................... | .................... |
| V | 7004 hCoV-19/Japan/PG-6794/2020\|EPI_ISL_687903\|2020-08 | .................... | .................... | ..................... | .................... | .................... | .................... |
| V | 7005 hCoV-19/Japan/PG-6795/2020\|EPI_ISL_687904\|2020-08 | .................... | .................... | ..................... | .................... | .................... | .................... |
| V | 7006 hCoV-19/Japan/PG-6796/2020\|EPI_ISL_687905\|2020-08 | .................... | .................... | ..................... | .................... | .................... | .................... |
| V | 7007 hCoV-19/Japan/PG-6797/2020\|EPI_ISL_687906\|2020-08 | .................... | .................... | ..................... | .................... | .................... | .................... |
| V | 7008 hCoV-19/Japan/PG-6798/2020\|EPI_ISL_687907\|2020-08 | .................... | .................... | ..................... | .................... | .................... | .................... |
| V | 7009 hCoV-19/Japan/PG-6799/2020\|EPI_ISL_687908\|2020-08 | .................... | .................... | ..................... | .................... | .................... | .................... |
| V | 7010 hCoV-19/Japan/PG-6800/2020\|EPI_ISL_687909\|2020-08 | .................... | .................... | ..................... | .................... | .................... | .................... |
| V | 7011 hCoV-19/Japan/PG-6801/2020\|EPI_ISL_687910\|2020-08 | .................... | .................... | ..................... | .................... | .................... | .................... |
| V | 7012 hCoV-19/Japan/PG-6802/2020\|EPI_ISL_687911\|2020-07 | .................... | .................... | ..................... | .................... | .................... | .................... |

| | | | | | | | | |
|---|---|---|---|---|---|---|---|---|
| V | 7013 | hCoV-19/Japan/PG-6803/2020\|EPI_ISL_687912\|2020-07 | .................... | .................... | .................... | .................... | .................... | .................... |
| V | 7014 | hCoV-19/Japan/PG-6804/2020\|EPI_ISL_687913\|2020-07 | .................... | .................... | .................... | .................... | .................... | .................... |
| V | 7015 | hCoV-19/Japan/PG-6805/2020\|EPI_ISL_687914\|2020-08 | .................... | .................... | .................... | .................... | .................... | .................... |
| V | 7016 | hCoV-19/Japan/PG-6806/2020\|EPI_ISL_687915\|2020-08 | .................... | .................... | .................... | .................... | .................... | .................... |
| V | 7017 | hCoV-19/Japan/PG-6807/2020\|EPI_ISL_687916\|2020-08 | .................... | .................... | .................... | .................... | .................... | .................... |
| V | 7018 | hCoV-19/Japan/PG-6808/2020\|EPI_ISL_687917\|2020-08 | .................... | .................... | .................... | .................... | .................... | .................... |
| V | 7019 | hCoV-19/Japan/PG-6809/2020\|EPI_ISL_687918\|2020-08 | .................... | .................... | .................... | .................... | .................... | .................... |
| V | 7020 | hCoV-19/Japan/PG-6810/2020\|EPI_ISL_687919\|2020-08 | .................... | .................... | .................... | .................... | .................... | .................... |
| V | 7021 | hCoV-19/Japan/PG-6811/2020\|EPI_ISL_687920\|2020-09 | .................... | .................... | .................... | .................... | .................... | .................... |
| V | 7022 | hCoV-19/Japan/PG-6812/2020\|EPI_ISL_687921\|2020-04 | .................... | .................... | .................... | .................... | .................... | .................... |
| V | 7023 | hCoV-19/Japan/PG-6813/2020\|EPI_ISL_687922\|2020-04 | .................... | .................... | .................... | .................... | .................... | .................... |
| V | 7024 | hCoV-19/Japan/PG-6814/2020\|EPI_ISL_687923\|2020-05 | .................... | .................... | .................... | .................... | .................... | .................... |
| V | 7025 | hCoV-19/Japan/PG-6815/2020\|EPI_ISL_687924\|2020-04 | .................... | .................... | .................... | .................... | .................... | .................... |
| V | 7026 | hCoV-19/Japan/PG-6816/2020\|EPI_ISL_687925\|2020-04 | .................... | .................... | .................... | .................... | .................... | .................... |
| V | 7027 | hCoV-19/Japan/PG-6818/2020\|EPI_ISL_687926\|2020-04 | .................... | .................... | .................... | .................... | .................... | .................... |
| V | 7028 | hCoV-19/Japan/PG-6819/2020\|EPI_ISL_687927\|2020-04 | .................... | .................... | .................... | .................... | .................... | .................... |
| V | 7029 | hCoV-19/Japan/PG-6820/2020\|EPI_ISL_687928\|2020-04 | .................... | .................... | .................... | .................... | .................... | .................... |
| V | 7030 | hCoV-19/Japan/PG-6821/2020\|EPI_ISL_687929\|2020-04 | .................... | .................... | .................... | .................... | .................... | .................... |
| V | 7031 | hCoV-19/Japan/PG-6822/2020\|EPI_ISL_687930\|2020-04 | .................... | .................... | .................... | .................... | .................... | .................... |
| V | 7032 | hCoV-19/Japan/PG-6823/2020\|EPI_ISL_687931\|2020-05 | .................... | .................... | .................... | .................... | .................... | .................... |
| V | 7033 | hCoV-19/Japan/PG-6824/2020\|EPI_ISL_687932\|2020-06 | .................... | .................... | .................... | .................... | .................... | .................... |
| V | 7034 | hCoV-19/Japan/PG-6825/2020\|EPI_ISL_687933\|2020-06 | .................... | .................... | .................... | .................... | .................... | .................... |
| V | 7035 | hCoV-19/Japan/PG-6826/2020\|EPI_ISL_687934\|2020-06 | .................... | .................... | .................... | .................... | .................... | .................... |
| V | 7036 | hCoV-19/Japan/PG-6827/2020\|EPI_ISL_687935\|2020-07 | .................... | .................... | .................... | .................... | .................... | .................... |
| V | 7037 | hCoV-19/Japan/PG-6828/2020\|EPI_ISL_687936\|2020-07 | .................... | .................... | .................... | .................... | .................... | .................... |
| V | 7038 | hCoV-19/Japan/PG-6829/2020\|EPI_ISL_687937\|2020-07 | .................... | .................... | .................... | .................... | .................... | .................... |
| V | 7039 | hCoV-19/Japan/PG-6830/2020\|EPI_ISL_687938\|2020-07 | .................... | .................... | .................... | .................... | .................... | .................... |
| V | 7040 | hCoV-19/Japan/PG-6831/2020\|EPI_ISL_687939\|2020-07 | .................... | .................... | .................... | .................... | .................... | .................... |
| V | 7041 | hCoV-19/Japan/PG-6832/2020\|EPI_ISL_687940\|2020-08 | .................... | .................... | .................... | .................... | .................... | .................... |
| V | 7042 | hCoV-19/Japan/PG-6833/2020\|EPI_ISL_687941\|2020-08 | .................... | .................... | .................... | .................... | .................... | .................... |
| V | 7043 | hCoV-19/Japan/PG-7378/2020\|EPI_ISL_688480\|2020-07 | .................... | .................... | .................... | .................... | .................... | .................... |
| V | 7044 | hCoV-19/Japan/PG-7379/2020\|EPI_ISL_688481\|2020-07 | .................... | .................... | .................... | .................... | .................... | .................... |
| V | 7045 | hCoV-19/Japan/PG-7380/2020\|EPI_ISL_688482\|2020-07 | .................... | .................... | .................... | .................... | .................... | .................... |
| V | 7046 | hCoV-19/Japan/PG-7381/2020\|EPI_ISL_688483\|2020-07 | .................... | .................... | .................... | .................... | .................... | .................... |
| V | 7047 | hCoV-19/Japan/PG-7382/2020\|EPI_ISL_688484\|2020-07 | .................... | .................... | .................... | .................... | .................... | .................... |
| V | 7048 | hCoV-19/Japan/PG-7383/2020\|EPI_ISL_688485\|2020-07 | .................... | .................... | .................... | .................... | .................... | .................... |
| V | 7049 | hCoV-19/Japan/PG-7384/2020\|EPI_ISL_688486\|2020-07 | .................... | .................... | .................... | .................... | .................... | .................... |
| V | 7050 | hCoV-19/Japan/PG-7385/2020\|EPI_ISL_688487\|2020-07 | .................... | .................... | .................... | .................... | .................... | .................... |
| V | 7051 | hCoV-19/Japan/PG-7386/2020\|EPI_ISL_688488\|2020-07 | .................... | .................... | .................... | .................... | .................... | .................... |
| V | 7052 | hCoV-19/Japan/PG-7387/2020\|EPI_ISL_688489\|2020-07 | .................... | .................... | .................... | .................... | .................... | .................... |
| V | 7053 | hCoV-19/Japan/PG-7388/2020\|EPI_ISL_688490\|2020-07 | .................... | .................... | .................... | .................... | .................... | .................... |
| V | 7054 | hCoV-19/Japan/PG-7389/2020\|EPI_ISL_688491\|2020-07 | .................... | .................... | .................... | .................... | .................... | .................... |
| V | 7055 | hCoV-19/Japan/PG-7390/2020\|EPI_ISL_688492\|2020-07 | .................... | .................... | .................... | .................... | .................... | .................... |
| V | 7056 | hCoV-19/Japan/PG-7391/2020\|EPI_ISL_688493\|2020-07 | .................... | .................... | .................... | .................... | .................... | .................... |
| V | 7057 | hCoV-19/Japan/PG-7392/2020\|EPI_ISL_688494\|2020-07 | .................... | .................... | .................... | .................... | .................... | .................... |
| V | 7058 | hCoV-19/Japan/PG-7393/2020\|EPI_ISL_688495\|2020-07 | .................... | .................... | .................... | .................... | .................... | .................... |
| V | 7059 | hCoV-19/Japan/PG-7394/2020\|EPI_ISL_688496\|2020-07 | .................... | .................... | .................... | .................... | .................... | .................... |
| V | 7060 | hCoV-19/Japan/PG-7395/2020\|EPI_ISL_688497\|2020-07 | .................... | .................... | .................... | .................... | .................... | .................... |
| V | 7061 | hCoV-19/Japan/PG-7396/2020\|EPI_ISL_688498\|2020-07 | .................... | .................... | .................... | .................... | .................... | .................... |
| V | 7062 | hCoV-19/Japan/PG-7397/2020\|EPI_ISL_688499\|2020-07 | .................... | .................... | .................... | .................... | .................... | .................... |
| V | 7063 | hCoV-19/Japan/PG-4614/2020\|EPI_ISL_689751\|2020-08 | .................... | .................... | .................... | .................... | .................... | .................... |
| V | 7064 | hCoV-19/Japan/PG-4615/2020\|EPI_ISL_689752\|2020-08 | .................... | .................... | .................... | .................... | .................... | .................... |
| V | 7065 | hCoV-19/Japan/PG-4616/2020\|EPI_ISL_689753\|2020-08 | .................... | .................... | .................... | .................... | .................... | .................... |
| V | 7066 | hCoV-19/Japan/PG-4617/2020\|EPI_ISL_689754\|2020-08 | .................... | .................... | .................... | .................... | .................... | .................... |
| V | 7067 | hCoV-19/Japan/PG-4618/2020\|EPI_ISL_689755\|2020-08 | .................... | .................... | .................... | .................... | .................... | .................... |
| V | 7068 | hCoV-19/Japan/PG-4619/2020\|EPI_ISL_689756\|2020-08 | .................... | .................... | .................... | .................... | .................... | .................... |
| V | 7069 | hCoV-19/Japan/PG-4620/2020\|EPI_ISL_689757\|2020-08 | .................... | .................... | .................... | .................... | .................... | .................... |
| V | 7070 | hCoV-19/Japan/PG-4621/2020\|EPI_ISL_689758\|2020-08 | .................... | .................... | .................... | .................... | .................... | .................... |
| V | 7071 | hCoV-19/Japan/PG-4622/2020\|EPI_ISL_689759\|2020-07 | .................... | .................... | .................... | .................... | .................... | .................... |
| V | 7072 | hCoV-19/Japan/PG-4623/2020\|EPI_ISL_689760\|2020-07 | .................... | .................... | .................... | .................... | .................... | .................... |
| V | 7073 | hCoV-19/Japan/PG-4624/2020\|EPI_ISL_689761\|2020-07 | .................... | .................... | .................... | .................... | .................... | .................... |
| V | 7074 | hCoV-19/Japan/PG-4625/2020\|EPI_ISL_689762\|2020-07 | .................... | .................... | .................... | .................... | .................... | .................... |
| V | 7075 | hCoV-19/Japan/PG-4626/2020\|EPI_ISL_689763\|2020-07 | .................... | .................... | .................... | .................... | .................... | .................... |
| V | 7076 | hCoV-19/Japan/PG-4627/2020\|EPI_ISL_689764\|2020-07 | .................... | .................... | .................... | .................... | .................... | .................... |
| V | 7077 | hCoV-19/Japan/PG-4628/2020\|EPI_ISL_689765\|2020-07 | .................... | .................... | .................... | .................... | .................... | .................... |
| V | 7078 | hCoV-19/Japan/PG-4629/2020\|EPI_ISL_689766\|2020-07 | .................... | .................... | .................... | .................... | .................... | .................... |
| V | 7079 | hCoV-19/Japan/PG-4630/2020\|EPI_ISL_689767\|2020-07 | .................... | .................... | .................... | .................... | .................... | .................... |
| V | 7080 | hCoV-19/Japan/PG-4631/2020\|EPI_ISL_689768\|2020-07 | .................... | .................... | .................... | .................... | ......T............. | .................... |
| V | 7081 | hCoV-19/Japan/PG-4632/2020\|EPI_ISL_689769\|2020-07 | .................... | .................... | .................... | .................... | .................... | .................... |
| V | 7082 | hCoV-19/Japan/PG-4633/2020\|EPI_ISL_689770\|2020-07 | .................... | .................... | .................... | .................... | .................... | .................... |
| V | 7083 | hCoV-19/Japan/PG-4634/2020\|EPI_ISL_689771\|2020-07 | .................... | .................... | .................... | .................... | .................... | .................... |
| V | 7084 | hCoV-19/Japan/PG-4635/2020\|EPI_ISL_689772\|2020-07 | .................... | .................... | .................... | .................... | .................... | .................... |
| V | 7085 | hCoV-19/Japan/PG-4636/2020\|EPI_ISL_689773\|2020-07 | .................... | .................... | .................... | .................... | .................... | .................... |
| V | 7086 | hCoV-19/Japan/PG-4637/2020\|EPI_ISL_689774\|2020-07 | .................... | .................... | .................... | .................... | .................... | .................... |
| V | 7087 | hCoV-19/Japan/PG-4638/2020\|EPI_ISL_689775\|2020-07 | .................... | .................... | .................... | .................... | .................... | .................... |
| V | 7088 | hCoV-19/Japan/PG-4639/2020\|EPI_ISL_689776\|2020-07 | .................... | .................... | .................... | .................... | .................... | .................... |
| V | 7089 | hCoV-19/Japan/PG-4640/2020\|EPI_ISL_689777\|2020-07 | .................... | .................... | .................... | .................... | .................... | .................... |
| V | 7090 | hCoV-19/Japan/PG-4641/2020\|EPI_ISL_689778\|2020-07 | .................... | .................... | .................... | .................... | .................... | .................... |
| V | 7091 | hCoV-19/Japan/PG-4642/2020\|EPI_ISL_689779\|2020-07 | .................... | .................... | .................... | .................... | .................... | .................... |
| V | 7092 | hCoV-19/Japan/PG-4643/2020\|EPI_ISL_689780\|2020-07 | .................... | .................... | .................... | .................... | .................... | .................... |

| | | | | | | | | |
|---|---|---|---|---|---|---|---|---|
| V | 7093 | hCoV-19/Japan/PG-4644/2020\|EPI_ISL_689781\|2020-07 | ···················· | ···················· | ···················· | ···················· | ···················· | ···················· |
| V | 7094 | hCoV-19/Japan/PG-4645/2020\|EPI_ISL_689782\|2020-07 | ···················· | ···················· | ···················· | ···················· | ···················· | ···················· |
| V | 7095 | hCoV-19/Japan/PG-4646/2020\|EPI_ISL_689783\|2020-07 | ···················· | ···················· | ···················· | ···················· | ···················· | ···················· |
| V | 7096 | hCoV-19/Japan/PG-4647/2020\|EPI_ISL_689784\|2020-07 | ···················· | ···················· | ···················· | ···················· | ···················· | ···················· |
| V | 7097 | hCoV-19/Japan/PG-4648/2020\|EPI_ISL_689785\|2020-07 | ···················· | ···················· | ···················· | ···················· | ···················· | ···················· |
| V | 7098 | hCoV-19/Japan/PG-4649/2020\|EPI_ISL_689786\|2020-07 | ···················· | ···················· | ···················· | ···················· | ···················· | ···················· |
| V | 7099 | hCoV-19/Japan/PG-4650/2020\|EPI_ISL_689787\|2020-07 | ···················· | ···················· | ···················· | ···················· | ···················· | ···················· |
| V | 7100 | hCoV-19/Japan/PG-4651/2020\|EPI_ISL_689788\|2020-07 | ···················· | ···················· | ···················· | ···················· | ···················· | ···················· |
| V | 7101 | hCoV-19/Japan/PG-4652/2020\|EPI_ISL_689789\|2020-07 | ···················· | ···················· | ···················· | ···················· | ···················· | ···················· |
| V | 7102 | hCoV-19/Japan/PG-4653/2020\|EPI_ISL_689790\|2020-07 | ···················· | ···················· | ···················· | ···················· | ···················· | ···················· |
| V | 7103 | hCoV-19/Japan/PG-4654/2020\|EPI_ISL_689791\|2020-07 | ···················· | ···················· | ···················· | ···················· | ···················· | ···················· |
| V | 7104 | hCoV-19/Japan/PG-4655/2020\|EPI_ISL_689792\|2020-07 | ···················· | ···················· | ···················· | ···················· | ···················· | ···················· |
| V | 7105 | hCoV-19/Japan/PG-4656/2020\|EPI_ISL_689793\|2020-07 | ···················· | ···················· | ···················· | ···················· | ···················· | ···················· |
| V | 7106 | hCoV-19/Japan/PG-4657/2020\|EPI_ISL_689794\|2020-07 | ···················· | ···················· | ···················· | ···················· | ···················· | ···················· |
| V | 7107 | hCoV-19/Japan/PG-4658/2020\|EPI_ISL_689795\|2020-08 | ···················· | ···················· | ···················· | ···················· | ···················· | ···················· |
| V | 7108 | hCoV-19/Japan/PG-4659/2020\|EPI_ISL_689796\|2020-08 | ···················· | ···················· | ···················· | ···················· | ···················· | ···················· |
| V | 7109 | hCoV-19/Japan/PG-4660/2020\|EPI_ISL_689797\|2020-08 | ···················· | ···················· | ···················· | ···················· | ···················· | ···················· |
| V | 7110 | hCoV-19/Japan/PG-4661/2020\|EPI_ISL_689798\|2020-08 | ···················· | ···················· | ···················· | ···················· | ···················· | ···················· |
| V | 7111 | hCoV-19/Japan/PG-4662/2020\|EPI_ISL_689799\|2020-08 | ···················· | ···················· | ···················· | ···················· | ···················· | ···················· |
| V | 7112 | hCoV-19/Japan/PG-2606/2020\|EPI_ISL_690740\|2020-04 | ···················· | ···················· | ···················· | ···················· | ···················· | ···················· |
| V | 7113 | hCoV-19/Japan/PG-2607/2020\|EPI_ISL_690741\|2020-05 | ···················· | ···················· | ···················· | ···················· | ···················· | ···················· |
| V | 7114 | hCoV-19/Japan/PG-2608/2020\|EPI_ISL_690742\|2020-05 | ···················· | ···················· | ···················· | ···················· | ···················· | ···················· |
| V | 7115 | hCoV-19/Japan/PG-2609/2020\|EPI_ISL_690743\|2020-05 | ···················· | ···················· | ···················· | ···················· | ···················· | ···················· |
| V | 7116 | hCoV-19/Japan/PG-2610/2020\|EPI_ISL_690744\|2020-06 | ···················· | ···················· | ···················· | ···················· | ···················· | ···················· |
| V | 7117 | hCoV-19/Japan/PG-2611/2020\|EPI_ISL_690745\|2020-02 | ···················· | ···················· | ···················· | ···················· | ···················· | ···················· |
| V | 7118 | hCoV-19/Japan/PG-2612/2020\|EPI_ISL_690746\|2020-03 | ···················· | ···················· | ···················· | ···················· | ···················· | ···················· |
| V | 7119 | hCoV-19/Japan/PG-2613/2020\|EPI_ISL_690747\|2020-03 | ···················· | ···················· | ···················· | ···················· | ···················· | ···················· |
| V | 7120 | hCoV-19/Japan/PG-2614/2020\|EPI_ISL_690748\|2020-03 | ···················· | ···················· | ···················· | ···················· | ···················· | ···················· |
| V | 7121 | hCoV-19/Japan/PG-2615/2020\|EPI_ISL_690749\|2020-03 | ···················· | ···················· | ···················· | ···················· | ···················· | ···················· |
| V | 7122 | hCoV-19/Japan/PG-2616/2020\|EPI_ISL_690750\|2020-03 | ···················· | ···················· | ···················· | ···················· | ···················· | ···················· |
| V | 7123 | hCoV-19/Japan/PG-2617/2020\|EPI_ISL_690751\|2020-03 | ···················· | ···················· | ···················· | ···················· | ···················· | ···················· |
| V | 7124 | hCoV-19/Japan/PG-2618/2020\|EPI_ISL_690752\|2020-04 | ···················· | ···················· | ···················· | ···················· | ···················· | ···················· |
| V | 7125 | hCoV-19/Japan/PG-2619/2020\|EPI_ISL_690753\|2020-04 | ···················· | ···················· | ···················· | ···················· | ···················· | ···················· |
| V | 7126 | hCoV-19/Japan/PG-2620/2020\|EPI_ISL_690754\|2020-04 | ···················· | ···················· | ···················· | ···················· | ···················· | ···················· |
| V | 7127 | hCoV-19/Japan/PG-2621/2020\|EPI_ISL_690755\|2020-04 | ···················· | ···················· | ···················· | ···················· | ···················· | ···················· |
| V | 7128 | hCoV-19/Japan/PG-2623/2020\|EPI_ISL_690756\|2020-04 | ···················· | ···················· | ···················· | ···················· | ···················· | ···················· |
| V | 7129 | hCoV-19/Japan/PG-2624/2020\|EPI_ISL_690757\|2020-04 | ···················· | ···················· | ···················· | ···················· | ···················· | ···················· |
| V | 7130 | hCoV-19/Japan/PG-2626/2020\|EPI_ISL_690758\|2020-04 | ···················· | ···················· | ···················· | ···················· | ···················· | ···················· |
| V | 7131 | hCoV-19/Japan/PG-2627/2020\|EPI_ISL_690759\|2020-04 | ···················· | ···················· | ···················· | ···················· | ···················· | ···················· |
| V | 7132 | hCoV-19/Japan/PG-2628/2020\|EPI_ISL_690760\|2020-04 | ···················· | ···················· | ···················· | ···················· | ···················· | ···················· |
| V | 7133 | hCoV-19/Japan/PG-2629/2020\|EPI_ISL_690761\|2020-04 | ···················· | ···················· | ···················· | ···················· | ···················· | ···················· |
| V | 7134 | hCoV-19/Japan/PG-2630/2020\|EPI_ISL_690762\|2020-04 | ···················· | ···················· | ···················· | ···················· | ···················· | ···················· |
| V | 7135 | hCoV-19/Japan/PG-2631/2020\|EPI_ISL_690763\|2020-04 | ···················· | ···················· | ···················· | ···················· | ···················· | ···················· |
| V | 7136 | hCoV-19/Japan/PG-2632/2020\|EPI_ISL_690764\|2020-04 | ···················· | ···················· | ···················· | ···················· | ···················· | ···················· |
| V | 7137 | hCoV-19/Japan/PG-2633/2020\|EPI_ISL_690765\|2020-04 | ···················· | ···················· | ···················· | ···················· | ···················· | ···················· |
| V | 7138 | hCoV-19/Japan/PG-2634/2020\|EPI_ISL_690766\|2020-04 | ···················· | ···················· | ···················· | ···················· | ···················· | ···················· |
| V | 7139 | hCoV-19/Japan/PG-2635/2020\|EPI_ISL_690767\|2020-04 | ···················· | ···················· | ···················· | ···················· | ···················· | ···················· |
| V | 7140 | hCoV-19/Japan/PG-2636/2020\|EPI_ISL_690768\|2020-04 | ···················· | ···················· | ···················· | ···················· | ···················· | ···················· |
| V | 7141 | hCoV-19/Japan/PG-2637/2020\|EPI_ISL_690769\|2020-04 | ···················· | ···················· | ···················· | ···················· | ···················· | ···················· |
| V | 7142 | hCoV-19/Japan/PG-2638/2020\|EPI_ISL_690770\|2020-04 | ···················· | ···················· | ···················· | ···················· | ···················· | ···················· |
| V | 7143 | hCoV-19/Japan/PG-2639/2020\|EPI_ISL_690771\|2020-04 | ···················· | ···················· | ···················· | ···················· | ···················· | ···················· |
| V | 7144 | hCoV-19/Japan/PG-2640/2020\|EPI_ISL_690772\|2020-04 | ···················· | ···················· | ···················· | ···················· | ···················· | ···················· |
| V | 7145 | hCoV-19/Japan/PG-2641/2020\|EPI_ISL_690773\|2020-04 | ···················· | ···················· | ···················· | ···················· | ···················· | ···················· |
| V | 7146 | hCoV-19/Japan/PG-2642/2020\|EPI_ISL_690774\|2020-04 | ···················· | ···················· | ···················· | ···················· | ···················· | ···················· |
| V | 7147 | hCoV-19/Japan/PG-2643/2020\|EPI_ISL_690775\|2020-04 | ···················· | ···················· | ···················· | ···················· | ···················· | ···················· |
| V | 7148 | hCoV-19/Japan/PG-2644/2020\|EPI_ISL_690776\|2020-04 | ···················· | ···················· | ···················· | ···················· | ···················· | ···················· |
| V | 7149 | hCoV-19/Japan/PG-2645/2020\|EPI_ISL_690777\|2020-04 | ···················· | ···················· | ···················· | ···················· | ···················· | ···················· |
| V | 7150 | hCoV-19/Japan/PG-2646/2020\|EPI_ISL_690778\|2020-06 | ···················· | ···················· | ···················· | ···················· | ···················· | ···················· |
| V | 7151 | hCoV-19/Japan/PG-2647/2020\|EPI_ISL_690779\|2020-04 | ···················· | ···················· | ···················· | ···················· | ···················· | ···················· |
| V | 7152 | hCoV-19/Japan/PG-2648/2020\|EPI_ISL_690780\|2020-04 | ···················· | ···················· | ···················· | ···················· | ···················· | ···················· |
| V | 7153 | hCoV-19/Japan/PG-2649/2020\|EPI_ISL_690781\|2020-04 | ···················· | ···················· | ···················· | ···················· | ···················· | ···················· |
| V | 7154 | hCoV-19/Japan/PG-2650/2020\|EPI_ISL_690782\|2020-04 | ···················· | ···················· | ···················· | ···················· | ···················· | ···················· |
| V | 7155 | hCoV-19/Japan/PG-2651/2020\|EPI_ISL_690783\|2020-04 | ···················· | ···················· | ···················· | ···················· | ···················· | ···················· |
| V | 7156 | hCoV-19/Japan/PG-2652/2020\|EPI_ISL_690784\|2020-04 | ···················· | ···················· | ···················· | ···················· | ···················· | ···················· |
| V | 7157 | hCoV-19/Japan/PG-2653/2020\|EPI_ISL_690785\|2020-04 | ···················· | ···················· | ···················· | ···················· | ···················· | ···················· |
| V | 7158 | hCoV-19/Japan/PG-2654/2020\|EPI_ISL_690786\|2020-04 | ···················· | ···················· | ···················· | ···················· | ···················· | ···················· |
| V | 7159 | hCoV-19/Japan/PG-2655/2020\|EPI_ISL_690787\|2020-04 | ···················· | ···················· | ···················· | ···················· | ···················· | ···················· |
| V | 7160 | hCoV-19/Japan/PG-2656/2020\|EPI_ISL_690788\|2020-04 | ···················· | ···················· | ···················· | ···················· | ···················· | ···················· |
| V | 7161 | hCoV-19/Japan/PG-2657/2020\|EPI_ISL_690789\|2020-04 | ···················· | ···················· | ···················· | ···················· | ···················· | ···················· |
| V | 7162 | hCoV-19/Japan/PG-2658/2020\|EPI_ISL_690790\|2020-04 | ···················· | ···················· | ···················· | ···················· | ···················· | ···················· |
| V | 7163 | hCoV-19/Japan/PG-2999/2020\|EPI_ISL_691123\|2020-04 | ···················· | ···················· | ···················· | ···················· | ···················· | ···················· |
| V | 7164 | hCoV-19/Japan/PG-3005/2020\|EPI_ISL_691129\|2020-04 | ···················· | ···················· | ···················· | ···················· | ···················· | ···················· |
| V | 7165 | hCoV-19/Japan/PG-4216/2020\|EPI_ISL_692454\|2020-07 | ···················· | ···················· | ···················· | ···················· | ···················· | ···················· |
| V | 7166 | hCoV-19/Japan/PG-4217/2020\|EPI_ISL_692455\|2020-07 | ···················· | ···················· | ···················· | ···················· | ···················· | ···················· |
| V | 7167 | hCoV-19/Switzerland/BL-ETHZ-350011/2020\|EPI_ISL_69 | ········T··········· | ···················· | ···················· | ···················· | ···················· | ···················· |
| V | 7168 | hCoV-19/Switzerland/GE-ETHZ-350012/2020\|EPI_ISL_69 | ···················· | ···················· | ···················· | ···················· | ···················· | ···················· |
| V | 7169 | hCoV-19/Japan/PG-1704/2020\|EPI_ISL_685310\|2020-04 | ···················· | ···················· | ···················· | ···················· | ···················· | ···················· |
| V | 7170 | hCoV-19/Japan/PG-1705/2020\|EPI_ISL_685311\|2020-05 | ···················· | ···················· | ···················· | ···················· | ···················· | ···················· |
| V | 7171 | hCoV-19/Japan/PG-1706/2020\|EPI_ISL_685312\|2020-05 | ···················· | ···················· | ···················· | ···················· | ···················· | ···················· |
| V | 7172 | hCoV-19/Japan/PG-1707/2020\|EPI_ISL_685313\|2020-04 | ···················· | ···················· | ···················· | ···················· | ···················· | ···················· |

| | | | | | | | | |
|---|---|---|---|---|---|---|---|---|
| V | 7173 | hCoV-19/Japan/PG-1708/2020\|EPI_ISL_685314\|2020-04 | .................... | .................... | .................... | .................... | .................... | .................... |
| V | 7174 | hCoV-19/Japan/PG-1709/2020\|EPI_ISL_685315\|2020-04 | .................... | ......A............. | .................... | .................... | .................... | .................... |
| V | 7175 | hCoV-19/Japan/PG-1710/2020\|EPI_ISL_685316\|2020-04 | .................... | .................... | .................... | .................... | .................... | .................... |
| V | 7176 | hCoV-19/Japan/PG-1711/2020\|EPI_ISL_685317\|2020-04 | .................... | .................... | .................... | .................... | .................... | .................... |
| V | 7177 | hCoV-19/Japan/PG-1712/2020\|EPI_ISL_685318\|2020-04 | .................... | .................... | .................... | .................... | .................... | .................... |
| V | 7178 | hCoV-19/Japan/PG-1713/2020\|EPI_ISL_685319\|2020-04 | .................... | .................... | .................... | .................... | .................... | .................... |
| V | 7179 | hCoV-19/Japan/PG-1714/2020\|EPI_ISL_685320\|2020-04 | .................... | .................... | .................... | .................... | .................... | .................... |
| V | 7180 | hCoV-19/Japan/PG-1715/2020\|EPI_ISL_685321\|2020-04 | .................... | .................... | .................... | .................... | .................... | .................... |
| V | 7181 | hCoV-19/Japan/PG-1716/2020\|EPI_ISL_685322\|2020-04 | .................... | .................... | .................... | .................... | .................... | .................... |
| V | 7182 | hCoV-19/Japan/PG-1717/2020\|EPI_ISL_685323\|2020-04 | .................... | .................... | .................... | .................... | .................... | .................... |
| V | 7183 | hCoV-19/Japan/PG-1718/2020\|EPI_ISL_685324\|2020-04 | .................... | .................... | .................... | .................... | .................... | .................... |
| V | 7184 | hCoV-19/Japan/PG-1719/2020\|EPI_ISL_685325\|2020-04 | .................... | .................... | .................... | .................... | .................... | .................... |
| V | 7185 | hCoV-19/Japan/PG-1720/2020\|EPI_ISL_685326\|2020-04 | .................... | .................... | .................... | .................... | .................... | .................... |
| V | 7186 | hCoV-19/Japan/PG-1721/2020\|EPI_ISL_685327\|2020-04 | .................... | .................... | .................... | .................... | .................... | .................... |
| V | 7187 | hCoV-19/Japan/PG-1722/2020\|EPI_ISL_685328\|2020-04 | .................... | .................... | .................... | .................... | .................... | .................... |
| V | 7188 | hCoV-19/Japan/PG-1723/2020\|EPI_ISL_685329\|2020-04 | .................... | .................... | .................... | .................... | .................... | .................... |
| V | 7189 | hCoV-19/Japan/PG-1724/2020\|EPI_ISL_685330\|2020-04 | .................... | .................... | .................... | .................... | .................... | .................... |
| V | 7190 | hCoV-19/Japan/PG-1725/2020\|EPI_ISL_685331\|2020-04 | .................... | .................... | .................... | .................... | .................... | .................... |
| V | 7191 | hCoV-19/Japan/PG-1726/2020\|EPI_ISL_685332\|2020-04 | .................... | .................... | .................... | .................... | .................... | .................... |
| V | 7192 | hCoV-19/Japan/PG-1727/2020\|EPI_ISL_685333\|2020-04 | .................... | .................... | .................... | .................... | .................... | .................... |
| V | 7193 | hCoV-19/Japan/PG-1728/2020\|EPI_ISL_685334\|2020-04 | .................... | .................... | .................... | .................... | .................... | .................... |
| V | 7194 | hCoV-19/Japan/PG-1729/2020\|EPI_ISL_685335\|2020-04 | .................... | .................... | .................... | .................... | .................... | .................... |
| V | 7195 | hCoV-19/Japan/PG-1730/2020\|EPI_ISL_685336\|2020-04 | .................... | .................... | .................... | .................... | .................... | .................... |
| V | 7196 | hCoV-19/Japan/PG-1731/2020\|EPI_ISL_685337\|2020-04 | .................... | .................... | .................... | .................... | .................... | .................... |
| V | 7197 | hCoV-19/Japan/PG-1732/2020\|EPI_ISL_685338\|2020-04 | .................... | ......A............. | .................... | .................... | .................... | .................... |
| V | 7198 | hCoV-19/Japan/PG-1733/2020\|EPI_ISL_685339\|2020-04 | .................... | .................... | .................... | .................... | .................... | .................... |
| V | 7199 | hCoV-19/Japan/PG-1734/2020\|EPI_ISL_685340\|2020-04 | .................... | .................... | .................... | .................... | .................... | .................... |
| V | 7200 | hCoV-19/Japan/PG-1735/2020\|EPI_ISL_685341\|2020-04 | .................... | .................... | .................... | .................... | .................... | .................... |
| V | 7201 | hCoV-19/Japan/PG-1736/2020\|EPI_ISL_685342\|2020-04 | .................... | .................... | .................... | .................... | .................... | .................... |
| V | 7202 | hCoV-19/Japan/PG-1737/2020\|EPI_ISL_685343\|2020-04 | .................... | .................... | .................... | .................... | .................... | .................... |
| V | 7203 | hCoV-19/Japan/PG-1738/2020\|EPI_ISL_685344\|2020-05 | .................... | .................... | .................... | .................... | .................... | .................... |
| V | 7204 | hCoV-19/Japan/PG-1739/2020\|EPI_ISL_685345\|2020-05 | .................... | .................... | .................... | .................... | .................... | .................... |
| V | 7205 | hCoV-19/Japan/PG-1740/2020\|EPI_ISL_685346\|2020-05 | .................... | .................... | .................... | .................... | .................... | .................... |
| V | 7206 | hCoV-19/Japan/PG-1741/2020\|EPI_ISL_685347\|2020-05 | .................... | ......A............. | .................... | .................... | .................... | .................... |
| V | 7207 | hCoV-19/Japan/PG-1742/2020\|EPI_ISL_685348\|2020-05 | .................... | .................... | .................... | .................... | .................... | .................... |
| V | 7208 | hCoV-19/Japan/PG-1743/2020\|EPI_ISL_685349\|2020-05 | .................... | .................... | .................... | .................... | .................... | .................... |
| V | 7209 | hCoV-19/Japan/PG-1744/2020\|EPI_ISL_685350\|2020-05 | .................... | ......A............. | .................... | .................... | .................... | .................... |
| V | 7210 | hCoV-19/Japan/PG-1745/2020\|EPI_ISL_685351\|2020-05 | .................... | ......A............. | .................... | .................... | .................... | .................... |
| V | 7211 | hCoV-19/Japan/PG-1746/2020\|EPI_ISL_685352\|2020-05 | .................... | .................... | .................... | .................... | .................... | .................... |
| V | 7212 | hCoV-19/Japan/PG-1747/2020\|EPI_ISL_685353\|2020-05 | .................... | .................... | .................... | .................... | .................... | .................... |
| V | 7213 | hCoV-19/Japan/PG-1748/2020\|EPI_ISL_685354\|2020-04 | .................... | .................... | .................... | .................... | .................... | .................... |
| V | 7214 | hCoV-19/Japan/PG-1749/2020\|EPI_ISL_685355\|2020-04 | .................... | .................... | .................... | .................... | .................... | .................... |
| V | 7215 | hCoV-19/Japan/PG-1750/2020\|EPI_ISL_685356\|2020-04 | .................... | .................... | .................... | .................... | .................... | .................... |
| V | 7216 | hCoV-19/Japan/PG-1751/2020\|EPI_ISL_685357\|2020-04 | .................... | .................... | .................... | .................... | .................... | .................... |
| V | 7217 | hCoV-19/Japan/PG-1752/2020\|EPI_ISL_685358\|2020-04 | .................... | .................... | .................... | .................... | .................... | .................... |
| V | 7218 | hCoV-19/Japan/PG-1753/2020\|EPI_ISL_685359\|2020-04 | .................... | .................... | .................... | .................... | .................... | .................... |
| V | 7219 | hCoV-19/Japan/PG-7537/2020\|EPI_ISL_686639\|2020-08 | .................... | .................... | .................... | .................... | .................... | .................... |
| V | 7220 | hCoV-19/Japan/PG-7538/2020\|EPI_ISL_686640\|2020-08 | .................... | .................... | .................... | .................... | .................... | .................... |
| V | 7221 | hCoV-19/Japan/PG-7539/2020\|EPI_ISL_686641\|2020-08 | .................... | .................... | .................... | .................... | .................... | .................... |
| V | 7222 | hCoV-19/Japan/PG-7540/2020\|EPI_ISL_686642\|2020-08 | .................... | .................... | .................... | .................... | .................... | .................... |
| V | 7223 | hCoV-19/Japan/PG-7541/2020\|EPI_ISL_686643\|2020-08 | .................... | .................... | .................... | .................... | .................... | .................... |
| V | 7224 | hCoV-19/Japan/PG-7542/2020\|EPI_ISL_686644\|2020-08 | .................... | .................... | .................... | .................... | .................... | .................... |
| V | 7225 | hCoV-19/Japan/PG-7543/2020\|EPI_ISL_686645\|2020-08 | .................... | .................... | .................... | .................... | .................... | .................... |
| V | 7226 | hCoV-19/Japan/PG-7544/2020\|EPI_ISL_686646\|2020-08 | .................... | .................... | .................... | .................... | .................... | .................... |
| V | 7227 | hCoV-19/Japan/PG-7545/2020\|EPI_ISL_686647\|2020-08 | .................... | .................... | .................... | .................... | .................... | .................... |
| V | 7228 | hCoV-19/Japan/PG-7546/2020\|EPI_ISL_686648\|2020-08 | .................... | .................... | .................... | .................... | .................... | .................... |
| V | 7229 | hCoV-19/Japan/PG-7547/2020\|EPI_ISL_686649\|2020-08 | .................... | .................... | .................... | .................... | .................... | .................... |
| V | 7230 | hCoV-19/Japan/PG-7548/2020\|EPI_ISL_686650\|2020-08 | .................... | .................... | .................... | .................... | .................... | .................... |
| V | 7231 | hCoV-19/Japan/PG-7549/2020\|EPI_ISL_686651\|2020-08 | .................... | .................... | .................... | .................... | .................... | .................... |
| V | 7232 | hCoV-19/Japan/PG-7550/2020\|EPI_ISL_686652\|2020-08 | .................... | .................... | .................... | .................... | .................... | .................... |
| V | 7233 | hCoV-19/Japan/PG-7551/2020\|EPI_ISL_686653\|2020-08 | .................... | .................... | .................... | .................... | .................... | .................... |
| V | 7235 | hCoV-19/Japan/PG-7553/2020\|EPI_ISL_686655\|2020-09 | .................... | .................... | .................... | .................... | .................... | .................... |
| V | 7236 | hCoV-19/Japan/PG-7554/2020\|EPI_ISL_686656\|2020-09 | .................... | .................... | .................... | .................... | .................... | .................... |
| V | 7237 | hCoV-19/Japan/PG-7555/2020\|EPI_ISL_686657\|2020-09 | .................... | .................... | .................... | .................... | .................... | .................... |
| V | 7238 | hCoV-19/Japan/PG-7556/2020\|EPI_ISL_686658\|2020-09 | .................... | .................... | .................... | .................... | .................... | .................... |
| V | 7239 | hCoV-19/Japan/PG-7557/2020\|EPI_ISL_686659\|2020-06 | .................... | .................... | .................... | .................... | .................... | .................... |
| V | 7240 | hCoV-19/Japan/PG-7558/2020\|EPI_ISL_686660\|2020-06 | .................... | .................... | .................... | .................... | .................... | .................... |
| V | 7241 | hCoV-19/Japan/PG-7559/2020\|EPI_ISL_686661\|2020-06 | .................... | .................... | .................... | .................... | .................... | .................... |
| V | 7242 | hCoV-19/Japan/PG-7560/2020\|EPI_ISL_686662\|2020-06 | .................... | .................... | .................... | .................... | .................... | .................... |
| V | 7243 | hCoV-19/Japan/PG-7561/2020\|EPI_ISL_686663\|2020-07 | .................... | .................... | .................... | .................... | .................... | .................... |
| V | 7244 | hCoV-19/Japan/PG-7562/2020\|EPI_ISL_686664\|2020-07 | .................... | .................... | .................... | .................... | .................... | .................... |
| V | 7245 | hCoV-19/Japan/PG-7563/2020\|EPI_ISL_686665\|2020-07 | .................... | .................... | .................... | .................... | .................... | .................... |
| V | 7246 | hCoV-19/Japan/PG-7564/2020\|EPI_ISL_686666\|2020-07 | .................... | .................... | .................... | .................... | .................... | .................... |
| V | 7247 | hCoV-19/Japan/PG-7565/2020\|EPI_ISL_686667\|2020-07 | .................... | .................... | .................... | .................... | .................... | .................... |
| V | 7248 | hCoV-19/Japan/PG-7566/2020\|EPI_ISL_686668\|2020-07 | .................... | .................... | .................... | .................... | .................... | .................... |
| V | 7249 | hCoV-19/Japan/PG-7567/2020\|EPI_ISL_686669\|2020-07 | .................... | .................... | .................... | .................... | .................... | .................... |
| V | 7250 | hCoV-19/Japan/PG-7568/2020\|EPI_ISL_686670\|2020-07 | .................... | .................... | .................... | .................... | .................... | .................... |
| V | 7251 | hCoV-19/Japan/PG-7569/2020\|EPI_ISL_686671\|2020-07 | .................... | .................... | .................... | .................... | .................... | .................... |
| V | 7252 | hCoV-19/Japan/PG-7570/2020\|EPI_ISL_686672\|2020-07 | .................... | .................... | .................... | .................... | .................... | .................... |
| V | 7253 | hCoV-19/Japan/PG-7571/2020\|EPI_ISL_686673\|2020-07 | .................... | .................... | .................... | .................... | .................... | .................... |

| | | | | | | | | |
|---|---|---|---|---|---|---|---|---|
| V | 7254 | hCoV-19/Japan/PG-7572/2020\|EPI_ISL_686674\|2020-07 | .................... | .................... | ..................... | .................... | .................... | .................... |
| V | 7255 | hCoV-19/Japan/PG-7573/2020\|EPI_ISL_686675\|2020-07 | .................... | .................... | ..................... | .................... | .................... | .................... |
| V | 7256 | hCoV-19/Japan/PG-7574/2020\|EPI_ISL_686676\|2020-07 | .................... | .................... | ..................... | .................... | .................... | .................... |
| V | 7257 | hCoV-19/Japan/PG-7575/2020\|EPI_ISL_686677\|2020-07 | .................... | .................... | ..................... | .................... | .................... | .................... |
| V | 7258 | hCoV-19/Japan/PG-7576/2020\|EPI_ISL_686678\|2020-07 | .................... | .................... | ..................... | .................... | .................... | .................... |
| V | 7259 | hCoV-19/Japan/PG-7577/2020\|EPI_ISL_686679\|2020-07 | .................... | .................... | ..................... | .................... | .................... | .................... |
| V | 7260 | hCoV-19/Japan/PG-7578/2020\|EPI_ISL_686680\|2020-07 | .................... | .................... | ..................... | .................... | .................... | .................... |
| V | 7261 | hCoV-19/Japan/PG-7579/2020\|EPI_ISL_686681\|2020-07 | .................... | .................... | ..................... | .................... | .................... | .................... |
| V | 7262 | hCoV-19/Japan/PG-7580/2020\|EPI_ISL_686682\|2020-07 | .................... | .................... | ..................... | .................... | .................... | .................... |
| V | 7263 | hCoV-19/Japan/PG-7581/2020\|EPI_ISL_686683\|2020-07 | .................... | .................... | ..................... | .................... | .................... | .................... |
| V | 7264 | hCoV-19/Japan/PG-7582/2020\|EPI_ISL_686684\|2020-07 | .................... | .................... | ..................... | .................... | .................... | .................... |
| V | 7265 | hCoV-19/Japan/PG-7583/2020\|EPI_ISL_686685\|2020-07 | .................... | .................... | ..................... | .................... | .................... | .................... |
| V | 7266 | hCoV-19/Japan/PG-7584/2020\|EPI_ISL_686686\|2020-07 | .................... | .................... | ..................... | .................... | .................... | .................... |
| V | 7267 | hCoV-19/Japan/PG-7585/2020\|EPI_ISL_686687\|2020-07 | .................... | .................... | ..................... | .................... | .................... | .................... |
| V | 7268 | hCoV-19/Japan/PG-7586/2020\|EPI_ISL_686688\|2020-07 | .................... | .................... | ..................... | .................... | .................... | .................... |
| V | 7269 | hCoV-19/Japan/PG-7587/2020\|EPI_ISL_686689\|2020-07 | .................... | .................... | ..................... | .................... | .................... | .................... |
| V | 7270 | hCoV-19/Japan/PG-7588/2020\|EPI_ISL_686690\|2020-07 | .................... | .................... | ..................... | .................... | .................... | .................... |
| V | 7271 | hCoV-19/Japan/PG-6862/2020\|EPI_ISL_687970\|2020-09 | .................... | .................... | ..................... | .................... | .................... | .................... |
| V | 7272 | hCoV-19/Japan/PG-6863/2020\|EPI_ISL_687971\|2020-09 | .................... | .................... | ..................... | .................... | .................... | .................... |
| V | 7273 | hCoV-19/Japan/PG-6864/2020\|EPI_ISL_687972\|2020-09 | .................... | .................... | ..................... | .................... | .................... | .................... |
| V | 7274 | hCoV-19/Japan/PG-6865/2020\|EPI_ISL_687973\|2020-06 | .................... | .................... | ..................... | .................... | .................... | .................... |
| V | 7275 | hCoV-19/Japan/PG-6866/2020\|EPI_ISL_687974\|2020-06 | .................... | .................... | ..................... | .................... | .................... | .................... |
| V | 7276 | hCoV-19/Japan/PG-6867/2020\|EPI_ISL_687975\|2020-06 | .................... | .................... | ..................... | .................... | .................... | .................... |
| V | 7277 | hCoV-19/Japan/PG-6868/2020\|EPI_ISL_687976\|2020-06 | .................... | .................... | ..................... | .................... | .................... | .................... |
| V | 7278 | hCoV-19/Japan/PG-6869/2020\|EPI_ISL_687977\|2020-06 | .................... | .................... | ..................... | .................... | .................... | .................... |
| V | 7279 | hCoV-19/Japan/PG-6870/2020\|EPI_ISL_687978\|2020-06 | .................... | .................... | ..................... | .................... | .................... | .................... |
| V | 7280 | hCoV-19/Japan/PG-6871/2020\|EPI_ISL_687979\|2020-06 | .................... | .................... | ..................... | .................... | .................... | .................... |
| V | 7281 | hCoV-19/Japan/PG-6872/2020\|EPI_ISL_687980\|2020-06 | .................... | .................... | ..................... | .................... | .................... | .................... |
| V | 7282 | hCoV-19/Japan/PG-6873/2020\|EPI_ISL_687981\|2020-06 | .................... | .................... | ..................... | .................... | .................... | .................... |
| V | 7283 | hCoV-19/Japan/PG-6874/2020\|EPI_ISL_687982\|2020-06 | .................... | .................... | ..................... | .................... | .................... | .................... |
| V | 7284 | hCoV-19/Japan/PG-6875/2020\|EPI_ISL_687983\|2020-06 | .................... | .................... | ..................... | .................... | .................... | .................... |
| V | 7285 | hCoV-19/Japan/PG-6876/2020\|EPI_ISL_687984\|2020-06 | .................... | .................... | ..................... | .................... | .................... | .................... |
| V | 7286 | hCoV-19/Japan/PG-6877/2020\|EPI_ISL_687985\|2020-06 | .................... | .................... | ..................... | .................... | .................... | .................... |
| V | 7287 | hCoV-19/Japan/PG-6878/2020\|EPI_ISL_687986\|2020-06 | .................... | .................... | ..................... | .................... | .................... | .................... |
| V | 7288 | hCoV-19/Japan/PG-6879/2020\|EPI_ISL_687987\|2020-06 | .................... | .................... | ..................... | .................... | .................... | .................... |
| V | 7289 | hCoV-19/Japan/PG-6880/2020\|EPI_ISL_687988\|2020-06 | .................... | .................... | ..................... | .................... | .................... | .................... |
| V | 7290 | hCoV-19/Japan/PG-6881/2020\|EPI_ISL_687989\|2020-06 | .................... | .................... | ..................... | .................... | .................... | .................... |
| V | 7291 | hCoV-19/Japan/PG-6882/2020\|EPI_ISL_687990\|2020-07 | .................... | .................... | ..................... | .................... | .................... | .................... |
| V | 7292 | hCoV-19/Japan/PG-6883/2020\|EPI_ISL_687991\|2020 | .................... | .................... | ..................... | .................... | .................... | .................... |
| V | 7293 | hCoV-19/Japan/PG-6884/2020\|EPI_ISL_687992\|2020 | .................... | .................... | ..................... | .................... | .................... | .................... |
| V | 7294 | hCoV-19/Japan/PG-6885/2020\|EPI_ISL_687993\|2020 | .................... | .................... | ..................... | .................... | .................... | .................... |
| V | 7295 | hCoV-19/Japan/PG-6886/2020\|EPI_ISL_687994\|2020 | .................... | .................... | ..................... | .................... | .................... | .................... |
| V | 7296 | hCoV-19/Japan/PG-6887/2020\|EPI_ISL_687995\|2020 | .................... | .................... | ..................... | .................... | .................... | .................... |
| V | 7297 | hCoV-19/Japan/PG-6888/2020\|EPI_ISL_687996\|2020 | .................... | .................... | ..................... | .................... | .................... | .................... |
| V | 7299 | hCoV-19/Japan/PG-6890/2020\|EPI_ISL_687998\|2020 | .................... | .................... | ..................... | .................... | .................... | .................... |
| V | 7300 | hCoV-19/Japan/PG-6891/2020\|EPI_ISL_687999\|2020 | .................... | .................... | ..................... | .................... | .................... | .................... |
| V | 7301 | hCoV-19/Japan/PG-2997/2020\|EPI_ISL_691121\|2020-04 | .................... | .................... | ..................... | .................... | .................... | .................... |
| V | 7302 | hCoV-19/Japan/PG-2998/2020\|EPI_ISL_691122\|2020-04 | .................... | .................... | ..................... | .................... | .................... | .................... |
| V | 7303 | hCoV-19/Japan/PG-3000/2020\|EPI_ISL_691124\|2020-04 | .................... | .................... | ..................... | .................... | .................... | .................... |
| V | 7304 | hCoV-19/Japan/PG-3001/2020\|EPI_ISL_691125\|2020-04 | .................... | .................... | ..................... | .................... | .................... | .................... |
| V | 7305 | hCoV-19/Japan/PG-3002/2020\|EPI_ISL_691126\|2020-04 | .................... | .................... | ..................... | .................... | .................... | .................... |
| V | 7306 | hCoV-19/Japan/PG-3003/2020\|EPI_ISL_691127\|2020-04 | .................... | .................... | ..................... | .................... | .................... | .................... |
| V | 7307 | hCoV-19/Japan/PG-3004/2020\|EPI_ISL_691128\|2020-04 | .................... | .................... | ..................... | .................... | .................... | .................... |
| V | 7308 | hCoV-19/Japan/PG-3006/2020\|EPI_ISL_691130\|2020-04 | .................... | .................... | ..................... | .................... | .................... | .................... |
| V | 7309 | hCoV-19/Japan/PG-3007/2020\|EPI_ISL_691131\|2020-04 | .................... | .................... | ..................... | .................... | .................... | .................... |
| V | 7310 | hCoV-19/Japan/PG-3008/2020\|EPI_ISL_691132\|2020-04 | .................... | .................... | ..................... | .................... | .................... | .................... |
| V | 7311 | hCoV-19/Japan/PG-3009/2020\|EPI_ISL_691133\|2020-04 | .................... | .................... | ..................... | .................... | .................... | .................... |
| V | 7312 | hCoV-19/Japan/PG-3010/2020\|EPI_ISL_691134\|2020-04 | .................... | .................... | ..................... | .................... | .................... | .................... |
| V | 7313 | hCoV-19/Japan/PG-3011/2020\|EPI_ISL_691135\|2020-04 | .................... | .................... | ..................... | .................... | .................... | .................... |
| V | 7314 | hCoV-19/Japan/PG-3012/2020\|EPI_ISL_691136\|2020-04 | .................... | .................... | ..................... | .................... | .................... | .................... |
| V | 7315 | hCoV-19/Japan/PG-3013/2020\|EPI_ISL_691137\|2020-04 | .................... | .................... | ..................... | .................... | .................... | .................... |
| V | 7317 | hCoV-19/Japan/PG-3015/2020\|EPI_ISL_691139\|2020-04 | .................... | .................... | ..................... | .................... | .................... | .................... |
| V | 7318 | hCoV-19/Japan/PG-3016/2020\|EPI_ISL_691140\|2020-04 | .................... | .................... | ..................... | .................... | .................... | .................... |
| V | 7319 | hCoV-19/Japan/PG-3017/2020\|EPI_ISL_691141\|2020-04 | .................... | .................... | ..................... | .................... | .................... | .................... |
| V | 7320 | hCoV-19/Japan/PG-3018/2020\|EPI_ISL_691142\|2020-04 | .................... | .................... | ..................... | .................... | .................... | .................... |
| V | 7321 | hCoV-19/Japan/PG-3019/2020\|EPI_ISL_691143\|2020-04 | .................... | .................... | ..................... | .................... | .................... | .................... |
| V | 7322 | hCoV-19/Japan/PG-3020/2020\|EPI_ISL_691144\|2020-04 | .................... | .................... | ..................... | .................... | .................... | .................... |
| V | 7323 | hCoV-19/Japan/PG-3021/2020\|EPI_ISL_691145\|2020-04 | .................... | .................... | ..................... | .................... | .................... | .................... |
| V | 7324 | hCoV-19/Japan/PG-3022/2020\|EPI_ISL_691146\|2020-04 | .................... | .................... | ..................... | .................... | .................... | .................... |
| V | 7325 | hCoV-19/Japan/PG-3023/2020\|EPI_ISL_691147\|2020-04 | .................... | .................... | ..................... | .................... | .................... | .................... |
| V | 7327 | hCoV-19/Japan/PG-3026/2020\|EPI_ISL_691149\|2020-04 | .................... | .................... | ..................... | .................... | .................... | .................... |
| V | 7328 | hCoV-19/Japan/PG-3027/2020\|EPI_ISL_691150\|2020-04 | .................... | .................... | ..................... | .................... | .................... | .................... |
| V | 7329 | hCoV-19/Japan/PG-3028/2020\|EPI_ISL_691151\|2020-04 | .................... | .................... | ..................... | .................... | .................... | .................... |
| V | 7330 | hCoV-19/Japan/PG-3029/2020\|EPI_ISL_691152\|2020-04 | .................... | .................... | ..................... | .................... | .................... | .................... |
| V | 7331 | hCoV-19/Japan/PG-3030/2020\|EPI_ISL_691153\|2020-04 | .................... | .................... | ..................... | .................... | .................... | .................... |
| V | 7332 | hCoV-19/Japan/PG-3031/2020\|EPI_ISL_691154\|2020-04 | .................... | .................... | ..................... | .................... | .................... | .................... |
| V | 7333 | hCoV-19/Japan/PG-3032/2020\|EPI_ISL_691155\|2020-04 | .................... | .................... | ..................... | .................... | .................... | .................... |
| V | 7334 | hCoV-19/Japan/PG-3033/2020\|EPI_ISL_691156\|2020-04 | .................... | .................... | ..................... | .................... | .................... | .................... |
| V | 7335 | hCoV-19/Japan/PG-3034/2020\|EPI_ISL_691157\|2020-04 | .................... | .................... | ..................... | .................... | .................... | .................... |
| V | 7336 | hCoV-19/Japan/PG-3035/2020\|EPI_ISL_691158\|2020-04 | .................... | .................... | ..................... | .................... | .................... | .................... |

| | | | | | | | | |
|---|---|---|---|---|---|---|---|---|
| V | 7337 | hCoV-19/Japan/PG-3036/2020\|EPI_ISL_691159\|2020-04 | ···················· | ···················· | ···················· | ···················· | ···················· | ···················· |
| V | 7338 | hCoV-19/Japan/PG-3037/2020\|EPI_ISL_691160\|2020-04 | ···················· | ···················· | ···················· | ···················· | ···················· | ···················· |
| V | 7339 | hCoV-19/Japan/PG-3038/2020\|EPI_ISL_691161\|2020-04 | ···················· | ···················· | ···················· | ···················· | ···················· | ···················· |
| V | 7340 | hCoV-19/Japan/PG-3039/2020\|EPI_ISL_691162\|2020-04 | ···················· | ···················· | ···················· | ···················· | ···················· | ···················· |
| V | 7341 | hCoV-19/Japan/PG-3040/2020\|EPI_ISL_691163\|2020-04 | ···················· | ···················· | ···················· | ···················· | ···················· | ···················· |
| V | 7342 | hCoV-19/Japan/PG-3041/2020\|EPI_ISL_691164\|2020-04 | ···················· | ···················· | ···················· | ···················· | ···················· | ···················· |
| V | 7343 | hCoV-19/Japan/PG-3042/2020\|EPI_ISL_691165\|2020-04 | ···················· | ···················· | ···················· | ···················· | ···················· | ···················· |
| V | 7344 | hCoV-19/Japan/PG-3043/2020\|EPI_ISL_691166\|2020-04 | ···················· | ···················· | ···················· | ···················· | ···················· | ···················· |
| V | 7345 | hCoV-19/Japan/PG-3044/2020\|EPI_ISL_691167\|2020-04 | ···················· | ···················· | ···················· | ···················· | ···················· | ···················· |
| V | 7346 | hCoV-19/Japan/PG-3045/2020\|EPI_ISL_691168\|2020-04 | ···················· | ···················· | ···················· | ···················· | ···················· | ···················· |
| V | 7347 | hCoV-19/Japan/PG-3046/2020\|EPI_ISL_691169\|2020-04 | ···················· | ···················· | ···················· | ···················· | ···················· | ···················· |
| V | 7348 | hCoV-19/Japan/PG-3047/2020\|EPI_ISL_691170\|2020-04 | ···················· | ···················· | ···················· | ···················· | ···················· | ···················· |
| V | 7349 | hCoV-19/Japan/PG-3048/2020\|EPI_ISL_691171\|2020-04 | ···················· | ···················· | ···················· | ···················· | ···················· | ···················· |
| V | 7350 | hCoV-19/Japan/PG-3049/2020\|EPI_ISL_691172\|2020-04 | ···················· | ···················· | ···················· | ···················· | ···················· | ···················· |
| V | 7351 | hCoV-19/Japan/PG-3050/2020\|EPI_ISL_691173\|2020-04 | ···················· | ···················· | ···················· | ···················· | ···················· | ···················· |
| V | 7352 | hCoV-19/Japan/PG-3051/2020\|EPI_ISL_691174\|2020-04 | ···················· | ···················· | ···················· | ···················· | ···················· | ···················· |
| V | 7353 | hCoV-19/Japan/PG-3052/2020\|EPI_ISL_691175\|2020-04 | ···················· | ···················· | ···················· | ···················· | ···················· | ···················· |
| V | 7354 | hCoV-19/Japan/PG-3053/2020\|EPI_ISL_691176\|2020-04 | ···················· | ···················· | ···················· | ···················· | ···················· | ···················· |
| V | 7355 | hCoV-19/Japan/PG-3054/2020\|EPI_ISL_691177\|2020-04 | ···················· | ···················· | ···················· | ···················· | ···················· | ···················· |
| V | 7356 | hCoV-19/Japan/PG-3055/2020\|EPI_ISL_691178\|2020-04 | ···················· | ···················· | ···················· | ···················· | ···················· | ···················· |
| V | 7357 | hCoV-19/Japan/PG-3056/2020\|EPI_ISL_691179\|2020-04 | ···················· | ···················· | ···················· | ···················· | ···················· | ···················· |
| V | 7358 | hCoV-19/Japan/PG-3057/2020\|EPI_ISL_691180\|2020-04 | ···················· | ···················· | ···················· | ···················· | ···················· | ···················· |
| V | 7359 | hCoV-19/Japan/PG-3058/2020\|EPI_ISL_691181\|2020-04 | ···················· | ···················· | ···················· | ···················· | ···················· | ···················· |
| V | 7360 | hCoV-19/Japan/PG-3059/2020\|EPI_ISL_691182\|2020-04 | ···················· | ···················· | ···················· | ···················· | ···················· | ···················· |
| V | 7361 | hCoV-19/Japan/PG-3060/2020\|EPI_ISL_691183\|2020-04 | ···················· | ···················· | ···················· | ···················· | ···················· | ···················· |
| V | 7362 | hCoV-19/Japan/PG-3061/2020\|EPI_ISL_691184\|2020-04 | ···················· | ···················· | ···················· | ···················· | ···················· | ···················· |
| V | 7363 | hCoV-19/Japan/PG-3062/2020\|EPI_ISL_691185\|2020-04 | ···················· | ···················· | ···················· | ···················· | ···················· | ···················· |
| V | 7364 | hCoV-19/Japan/PG-3063/2020\|EPI_ISL_691186\|2020-04 | ···················· | ···················· | ···················· | ···················· | ···················· | ···················· |
| V | 7365 | hCoV-19/Japan/PG-4214/2020\|EPI_ISL_692452\|2020-07 | ···················· | ···················· | ···················· | ···················· | ···················· | ···················· |
| V | 7366 | hCoV-19/Japan/PG-4215/2020\|EPI_ISL_692453\|2020-07 | ···················· | ···················· | ···················· | ···················· | ···················· | ···················· |
| V | 7367 | hCoV-19/Japan/PG-4218/2020\|EPI_ISL_692456\|2020-07 | ···················· | ···················· | ···················· | ···················· | ···················· | ···················· |
| V | 7368 | hCoV-19/Japan/PG-4219/2020\|EPI_ISL_692457\|2020-07 | ···················· | ···················· | ···················· | ···················· | ···················· | ···················· |
| V | 7369 | hCoV-19/Japan/PG-4220/2020\|EPI_ISL_692458\|2020-07 | ···················· | ···················· | ···················· | ···················· | ···················· | ···················· |
| V | 7370 | hCoV-19/Japan/PG-4221/2020\|EPI_ISL_692459\|2020-07 | ···················· | ···················· | ···················· | ···················· | ···················· | ···················· |
| V | 7371 | hCoV-19/Japan/PG-4222/2020\|EPI_ISL_692460\|2020-07 | ···················· | ···················· | ···················· | ···················· | ···················· | ···················· |
| V | 7372 | hCoV-19/Japan/PG-4223/2020\|EPI_ISL_692461\|2020-07 | ···················· | ···················· | ···················· | ···················· | ···················· | ···················· |
| V | 7373 | hCoV-19/Japan/PG-4224/2020\|EPI_ISL_692462\|2020-07 | ···················· | ···················· | ···················· | ···················· | ···················· | ···················· |
| V | 7374 | hCoV-19/Japan/PG-4225/2020\|EPI_ISL_692463\|2020-07 | ···················· | ···················· | ···················· | ···················· | ···················· | ···················· |
| V | 7375 | hCoV-19/Japan/PG-4226/2020\|EPI_ISL_692464\|2020-07 | ···················· | ···················· | ···················· | ···················· | ···················· | ···················· |
| V | 7376 | hCoV-19/Japan/PG-4227/2020\|EPI_ISL_692465\|2020-07 | ···················· | ···················· | ···················· | ···················· | ···················· | ···················· |
| V | 7377 | hCoV-19/Japan/PG-4228/2020\|EPI_ISL_692466\|2020-07 | ···················· | ···················· | ···················· | ···················· | ···················· | ···················· |
| V | 7378 | hCoV-19/Japan/PG-4230/2020\|EPI_ISL_692467\|2020-07 | ···················· | ···················· | ···················· | ···················· | ···················· | ···················· |
| V | 7379 | hCoV-19/Japan/PG-4231/2020\|EPI_ISL_692468\|2020-07 | ···················· | ···················· | ···················· | ···················· | ···················· | ···················· |
| V | 7380 | hCoV-19/Japan/PG-4232/2020\|EPI_ISL_692469\|2020-07 | ···················· | ···················· | ···················· | ···················· | ···················· | ···················· |
| V | 7381 | hCoV-19/Japan/PG-4240/2020\|EPI_ISL_692470\|2020-07 | ···················· | ···················· | ···················· | ···················· | ···················· | ···················· |
| V | 7382 | hCoV-19/Japan/PG-4241/2020\|EPI_ISL_692471\|2020-07 | ···················· | ···················· | ···················· | ···················· | ···················· | ···················· |
| V | 7383 | hCoV-19/Japan/PG-4242/2020\|EPI_ISL_692472\|2020-07 | ···················· | ···················· | ···················· | ···················· | ···················· | ···················· |
| V | 7384 | hCoV-19/Japan/PG-4243/2020\|EPI_ISL_692473\|2020-07 | ···················· | ···················· | ···················· | ···················· | ···················· | ···················· |
| V | 7385 | hCoV-19/Japan/PG-4244/2020\|EPI_ISL_692474\|2020-07 | ···················· | ···················· | ···················· | ···················· | ···················· | ···················· |
| V | 7386 | hCoV-19/Japan/PG-4245/2020\|EPI_ISL_692475\|2020-07 | ···················· | ···················· | ···················· | ···················· | ···················· | ···················· |
| V | 7387 | hCoV-19/Japan/PG-4246/2020\|EPI_ISL_692476\|2020-07 | ···················· | ···················· | ···················· | ···················· | ···················· | ···················· |
| V | 7388 | hCoV-19/Japan/PG-4247/2020\|EPI_ISL_692477\|2020-07 | ···················· | ···················· | ···················· | ···················· | ···················· | ···················· |
| V | 7389 | hCoV-19/Japan/PG-4248/2020\|EPI_ISL_692478\|2020-07 | ···················· | ···················· | ···················· | ···················· | ···················· | ···················· |
| V | 7390 | hCoV-19/Japan/PG-4249/2020\|EPI_ISL_692479\|2020-07 | ···················· | ···················· | ···················· | ···················· | ···················· | ···················· |
| V | 7391 | hCoV-19/Japan/PG-4250/2020\|EPI_ISL_692480\|2020-07 | ···················· | ···················· | ···················· | ···················· | ···················· | ···················· |
| V | 7392 | hCoV-19/Japan/PG-4251/2020\|EPI_ISL_692481\|2020-07 | ···················· | ···················· | ···················· | ···················· | ···················· | ···················· |
| V | 7393 | hCoV-19/Japan/PG-4252/2020\|EPI_ISL_692482\|2020-07 | ···················· | ···················· | ···················· | ···················· | ···················· | ···················· |
| V | 7394 | hCoV-19/Japan/PG-4253/2020\|EPI_ISL_692483\|2020-07 | ···················· | ···················· | ···················· | ···················· | ···················· | ···················· |
| V | 7395 | hCoV-19/Japan/PG-4254/2020\|EPI_ISL_692484\|2020-07 | ···················· | ···················· | ···················· | ···················· | ···················· | ···················· |
| V | 7396 | hCoV-19/Japan/PG-4255/2020\|EPI_ISL_692485\|2020-07 | ···················· | ···················· | ···················· | ···················· | ···················· | ···················· |
| V | 7397 | hCoV-19/Japan/PG-4256/2020\|EPI_ISL_692486\|2020-07 | ···················· | ···················· | ···················· | ···················· | ···················· | ···················· |
| V | 7398 | hCoV-19/Japan/PG-4257/2020\|EPI_ISL_692487\|2020-07 | ···················· | ···················· | ···················· | ···················· | ···················· | ···················· |
| V | 7399 | hCoV-19/Japan/PG-4258/2020\|EPI_ISL_692488\|2020-07 | ···················· | ···················· | ···················· | ···················· | ···················· | ···················· |
| V | 7400 | hCoV-19/Japan/PG-4259/2020\|EPI_ISL_692489\|2020-07 | ···················· | ···················· | ···················· | ···················· | ···················· | ···················· |
| V | 7401 | hCoV-19/Japan/PG-4260/2020\|EPI_ISL_692490\|2020-07 | ···················· | ···················· | ···················· | ···················· | ···················· | ···················· |
| V | 7402 | hCoV-19/Japan/PG-4261/2020\|EPI_ISL_692491\|2020-07 | ···················· | ···················· | ···················· | ···················· | ···················· | ···················· |
| V | 7403 | hCoV-19/Japan/PG-4262/2020\|EPI_ISL_692492\|2020-07 | ···················· | ···················· | ···················· | ···················· | ···················· | ···················· |
| V | 7404 | hCoV-19/Japan/PG-4263/2020\|EPI_ISL_692493\|2020-07 | ···················· | ···················· | ···················· | ···················· | ···················· | ···················· |
| V | 7405 | hCoV-19/Japan/PG-4264/2020\|EPI_ISL_692494\|2020-07 | ···················· | ···················· | ···················· | ···················· | ···················· | ···················· |
| V | 7406 | hCoV-19/Japan/PG-4265/2020\|EPI_ISL_692495\|2020-07 | ···················· | ···················· | ···················· | ···················· | ···················· | ···················· |
| V | 7407 | hCoV-19/Japan/PG-4266/2020\|EPI_ISL_692496\|2020-07 | ···················· | ···················· | ···················· | ···················· | ···················· | ···················· |
| V | 7408 | hCoV-19/Japan/PG-4267/2020\|EPI_ISL_692497\|2020-07 | ···················· | ···················· | ···················· | ···················· | ···················· | ···················· |
| V | 7409 | hCoV-19/Japan/PG-4268/2020\|EPI_ISL_692498\|2020-07 | ···················· | ···················· | ···················· | ···················· | ···················· | ···················· |
| V | 7410 | hCoV-19/Japan/PG-4269/2020\|EPI_ISL_692499\|2020-07 | ···················· | ···················· | ···················· | ···················· | ···················· | ···················· |
| V | 7411 | hCoV-19/Switzerland/BL-ETHZ-350009/2020\|EPI_ISL_69 | ···················· | ···················· | ···················· | ···················· | ···················· | ···················· |
| V | 7412 | hCoV-19/Switzerland/BL-ETHZ-350010/2020\|EPI_ISL_69 | ···················· | ···················· | ···················· | ···················· | ···················· | ···················· |
| V | 7413 | hCoV-19/Switzerland/BL-ETHZ-350013/2020\|EPI_ISL_69 | ···················· | ···················· | ···················· | ···················· | ···················· | ···················· |
| V | 7414 | hCoV-19/Switzerland/BL-ETHZ-350014/2020\|EPI_ISL_69 | ···················· | ···················· | ···················· | ···················· | ······K············· | ···················· |
| V | 7415 | hCoV-19/Switzerland/JU-ETHZ-350015/2020\|EPI_ISL_69 | ···················· | ···················· | ···················· | ···················· | ···················· | ···················· |
| V | 7416 | hCoV-19/Switzerland/TI-ETHZ-350016/2020\|EPI_ISL_69 | ···················· | ···················· | ···················· | ···················· | ···················· | ···················· |

| | | | | | | | |
|---|---|---|---|---|---|---|---|
| V | 7417 hCoV-19/Switzerland/VS-ETHZ-350017/2020\|EPI_ISL_69 | .................... | .................... | ..................... | .................... | .................... | .................... |
| V | 7418 hCoV-19/Switzerland/ZH-ETHZ-350018/2020\|EPI_ISL_69 | .................... | .................... | ..................... | .................... | .................... | .................... |
| V | 7419 hCoV-19/Switzerland/BL-ETHZ-350019/2020\|EPI_ISL_69 | .................... | .................... | ..................... | .................... | .................... | .................... |
| V | 7420 hCoV-19/Switzerland/SO-ETHZ-350094/2020\|EPI_ISL_69 | .................... | .................... | ..................... | .................... | .................... | .................... |
| V | 7421 hCoV-19/Switzerland/GE-ETHZ-350095/2020\|EPI_ISL_69 | .................... | .................... | ..................... | .................... | .................... | .................... |
| V | 7422 hCoV-19/Switzerland/BE-ETHZ-350096/2020\|EPI_ISL_69 | .................... | .................... | ..................... | .................... | .................... | .................... |
| V | 7423 hCoV-19/Switzerland/BE-ETHZ-350098/2020\|EPI_ISL_69 | .................... | .................... | ..................... | .................... | .................... | .................... |
| V | 7424 hCoV-19/Switzerland/VS-ETHZ-350099/2020\|EPI_ISL_69 | .................... | .................... | ..................... | .................... | .................... | .................... |
| V | 7425 hCoV-19/Japan/PG-1644/2020\|EPI_ISL_685250\|2020-04 | .................... | .................... | ..................... | .................... | .................... | .................... |
| V | 7426 hCoV-19/Japan/PG-1645/2020\|EPI_ISL_685251\|2020-04 | .................... | .................... | ..................... | .................... | .................... | .................... |
| V | 7427 hCoV-19/Japan/PG-1646/2020\|EPI_ISL_685252\|2020-04 | .................... | .................... | ..................... | .................... | .................... | .................... |
| V | 7428 hCoV-19/Japan/PG-1647/2020\|EPI_ISL_685253\|2020-04 | .................... | .................... | ..................... | .................... | .................... | .................... |
| V | 7429 hCoV-19/Japan/PG-1648/2020\|EPI_ISL_685254\|2020-04 | .................... | .................... | ..................... | .................... | .................... | .................... |
| V | 7430 hCoV-19/Japan/PG-1649/2020\|EPI_ISL_685255\|2020-04 | .................... | .................... | ..................... | .................... | .................... | .................... |
| V | 7431 hCoV-19/Japan/PG-1650/2020\|EPI_ISL_685256\|2020-04 | .................... | .................... | ..................... | .................... | .................... | .................... |
| V | 7432 hCoV-19/Japan/PG-1651/2020\|EPI_ISL_685257\|2020-04 | .................... | .................... | ..................... | .................... | .................... | .................... |
| V | 7433 hCoV-19/Japan/PG-1652/2020\|EPI_ISL_685258\|2020-04 | .................... | .................... | ..................... | .................... | .................... | .................... |
| V | 7434 hCoV-19/Japan/PG-1653/2020\|EPI_ISL_685259\|2020-04 | .................... | .................... | ..................... | .................... | .................... | .................... |
| V | 7435 hCoV-19/Japan/PG-1654/2020\|EPI_ISL_685260\|2020-04 | .................... | .................... | ..................... | .................... | .................... | .................... |
| V | 7436 hCoV-19/Japan/PG-1655/2020\|EPI_ISL_685261\|2020-04 | .................... | .................... | ..................... | .................... | .................... | .................... |
| V | 7437 hCoV-19/Japan/PG-1656/2020\|EPI_ISL_685262\|2020-04 | .................... | .................... | ..................... | .................... | .................... | .................... |
| V | 7438 hCoV-19/Japan/PG-1657/2020\|EPI_ISL_685263\|2020-04 | .................... | .................... | ..................... | .................... | .................... | .................... |
| V | 7439 hCoV-19/Japan/PG-1658/2020\|EPI_ISL_685264\|2020-04 | .................... | .................... | ..................... | .................... | .................... | .................... |
| V | 7440 hCoV-19/Japan/PG-1659/2020\|EPI_ISL_685265\|2020-04 | .................... | .................... | ..................... | .................... | .................... | .................... |
| V | 7441 hCoV-19/Japan/PG-1660/2020\|EPI_ISL_685266\|2020-04 | .................... | .................... | ..................... | .................... | .................... | .................... |
| V | 7442 hCoV-19/Japan/PG-1661/2020\|EPI_ISL_685267\|2020-04 | .................... | .................... | ..................... | .................... | .................... | .................... |
| V | 7443 hCoV-19/Japan/PG-1662/2020\|EPI_ISL_685268\|2020-04 | .................... | .................... | ..................... | .................... | .................... | .................... |
| V | 7444 hCoV-19/Japan/PG-1663/2020\|EPI_ISL_685269\|2020-04 | .................... | .................... | ..................... | .................... | .................... | .................... |
| V | 7445 hCoV-19/Japan/PG-1664/2020\|EPI_ISL_685270\|2020-04 | .................... | .................... | ..................... | .................... | .................... | .................... |
| V | 7446 hCoV-19/Japan/PG-1665/2020\|EPI_ISL_685271\|2020-04 | .................... | .................... | ..................... | .................... | .................... | .................... |
| V | 7447 hCoV-19/Japan/PG-1666/2020\|EPI_ISL_685272\|2020-04 | .................... | .................... | ..................... | .................... | .................... | .................... |
| V | 7448 hCoV-19/Japan/PG-1667/2020\|EPI_ISL_685273\|2020-04 | .................... | .................... | ..................... | .................... | .................... | .................... |
| V | 7449 hCoV-19/Japan/PG-1668/2020\|EPI_ISL_685274\|2020-04 | .................... | .................... | ..................... | .................... | .................... | .................... |
| V | 7450 hCoV-19/Japan/PG-1669/2020\|EPI_ISL_685275\|2020-04 | .................... | .................... | ..................... | .................... | .................... | .................... |
| V | 7451 hCoV-19/Japan/PG-1670/2020\|EPI_ISL_685276\|2020-04 | .................... | .................... | ..................... | .................... | .................... | .................... |
| V | 7452 hCoV-19/Japan/PG-1671/2020\|EPI_ISL_685277\|2020-04 | .................... | .................... | ..................... | .................... | .................... | .................... |
| V | 7453 hCoV-19/Japan/PG-1672/2020\|EPI_ISL_685278\|2020-04 | .................... | .................... | ..................... | .................... | .................... | .................... |
| V | 7454 hCoV-19/Japan/PG-1673/2020\|EPI_ISL_685279\|2020-04 | .................... | .................... | ..................... | .................... | .................... | .................... |
| V | 7455 hCoV-19/Japan/PG-1674/2020\|EPI_ISL_685280\|2020-04 | .................... | .................... | ..................... | .................... | .................... | .................... |
| V | 7456 hCoV-19/Japan/PG-1675/2020\|EPI_ISL_685281\|2020-05 | .................... | .................... | ..................... | .................... | .................... | .................... |
| V | 7457 hCoV-19/Japan/PG-1676/2020\|EPI_ISL_685282\|2020-04 | .................... | .................... | ..................... | .................... | .................... | .................... |
| V | 7458 hCoV-19/Japan/PG-1677/2020\|EPI_ISL_685283\|2020-05 | .................... | .................... | ..................... | .................... | .................... | .................... |
| V | 7459 hCoV-19/Japan/PG-1678/2020\|EPI_ISL_685284\|2020-05 | .................... | .................... | ..................... | .................... | .................... | .................... |
| V | 7460 hCoV-19/Japan/PG-1679/2020\|EPI_ISL_685285\|2020-05 | .................... | .................... | ..................... | .................... | .................... | .................... |
| V | 7461 hCoV-19/Japan/PG-1680/2020\|EPI_ISL_685286\|2020-05 | .................... | .................... | ..................... | .................... | .................... | .................... |
| V | 7462 hCoV-19/Japan/PG-1681/2020\|EPI_ISL_685287\|2020-05 | .................... | .................... | ..................... | .................... | .................... | .................... |
| V | 7463 hCoV-19/Japan/PG-1682/2020\|EPI_ISL_685288\|2020-05 | .................... | .................... | ..................... | .................... | .................... | .................... |
| V | 7464 hCoV-19/Japan/PG-1683/2020\|EPI_ISL_685289\|2020-05 | .................... | .................... | ..................... | .................... | .................... | .................... |
| V | 7465 hCoV-19/Japan/PG-1684/2020\|EPI_ISL_685290\|2020-05 | .................... | .................... | ..................... | .................... | .................... | .................... |
| V | 7466 hCoV-19/Japan/PG-1685/2020\|EPI_ISL_685291\|2020-05 | .................... | .................... | ..................... | .................... | .................... | .................... |
| V | 7467 hCoV-19/Japan/PG-1686/2020\|EPI_ISL_685292\|2020-05 | .................... | .................... | ..................... | .................... | .................... | .................... |
| V | 7468 hCoV-19/Japan/PG-1687/2020\|EPI_ISL_685293\|2020-05 | .................... | .................... | ..................... | .................... | .................... | .................... |
| V | 7469 hCoV-19/Japan/PG-1688/2020\|EPI_ISL_685294\|2020-05 | .................... | .................... | ..................... | .................... | .................... | .................... |
| V | 7470 hCoV-19/Japan/PG-1689/2020\|EPI_ISL_685295\|2020-05 | .................... | .................... | ..................... | .................... | .................... | .................... |
| V | 7471 hCoV-19/Japan/PG-1690/2020\|EPI_ISL_685296\|2020-05 | .................... | .................... | ..................... | .................... | .................... | .................... |
| V | 7472 hCoV-19/Japan/PG-1691/2020\|EPI_ISL_685297\|2020-03 | .................... | .................... | ..................... | .................... | .................... | .................... |
| V | 7473 hCoV-19/Japan/PG-1692/2020\|EPI_ISL_685298\|2020-03 | .................... | .................... | ..................... | .................... | .................... | .................... |
| V | 7474 hCoV-19/Japan/PG-1693/2020\|EPI_ISL_685299\|2020-04 | .................... | .................... | ..................... | .................... | .................... | .................... |
| V | 7475 hCoV-19/Japan/PG-1694/2020\|EPI_ISL_685300\|2020-04 | .................... | .................... | ..................... | .................... | .................... | .................... |
| V | 7476 hCoV-19/Japan/PG-1695/2020\|EPI_ISL_685301\|2020-04 | .................... | .................... | ..................... | .................... | .................... | .................... |
| V | 7477 hCoV-19/Japan/PG-1696/2020\|EPI_ISL_685302\|2020-04 | .................... | .................... | ..................... | .................... | .................... | .................... |
| V | 7478 hCoV-19/Japan/PG-1697/2020\|EPI_ISL_685303\|2020-04 | .................... | .................... | ..................... | .................... | .................... | .................... |
| V | 7479 hCoV-19/Japan/PG-1698/2020\|EPI_ISL_685304\|2020-04 | .................... | .................... | ..................... | .................... | .................... | .................... |
| V | 7480 hCoV-19/Japan/PG-1699/2020\|EPI_ISL_685305\|2020-04 | .................... | .................... | ..................... | .................... | .................... | .................... |
| V | 7481 hCoV-19/Japan/PG-1700/2020\|EPI_ISL_685306\|2020-04 | .................... | .................... | ..................... | .................... | .................... | .................... |
| V | 7482 hCoV-19/Japan/PG-1701/2020\|EPI_ISL_685307\|2020-04 | .................... | .................... | ..................... | .................... | .................... | .................... |
| V | 7483 hCoV-19/Japan/PG-1702/2020\|EPI_ISL_685308\|2020-04 | .................... | .................... | ..................... | .................... | .................... | .................... |
| V | 7484 hCoV-19/Japan/PG-1703/2020\|EPI_ISL_685309\|2020-04 | .................... | .................... | ..................... | .................... | .................... | .................... |
| V | 7485 hCoV-19/England/204661791/2020\|EPI_ISL_686581\|2020 | .................... | .................... | ..................... | .................... | .................... | .................... |
| V | 7486 hCoV-19/England/204661792/2020\|EPI_ISL_686582\|2020 | .................... | .................... | ..................... | .................... | .................... | .................... |
| V | 7487 hCoV-19/England/204681084/2020\|EPI_ISL_686583\|2020 | .................... | .................... | ..................... | .................... | .................... | .................... |
| V | 7488 hCoV-19/England/204690804/2020\|EPI_ISL_686584\|2020 | .................... | .................... | ..................... | .................... | .................... | .................... |
| V | 7489 hCoV-19/England/204690817/2020\|EPI_ISL_686585\|2020 | .................... | .................... | ..................... | .................... | .................... | .................... |
| V | 7490 hCoV-19/England/204690835/2020\|EPI_ISL_686586\|2020 | .................... | .................... | ..................... | .................... | .................... | .................... |
| V | 7491 hCoV-19/England/204690838/2020\|EPI_ISL_686587\|2020 | .................... | .................... | ..................... | .................... | .................... | .................... |
| V | 7492 hCoV-19/England/204690872/2020\|EPI_ISL_686588\|2020 | .................... | .................... | ..................... | .................... | .................... | .................... |
| V | 7493 hCoV-19/England/204690892/2020\|EPI_ISL_686589\|2020 | .................... | .................... | ..................... | .................... | .................... | .................... |
| V | 7494 hCoV-19/England/204690898/2020\|EPI_ISL_686590\|2020 | .................... | .................... | ..................... | .................... | .................... | .................... |
| V | 7495 hCoV-19/England/204690958/2020\|EPI_ISL_686591\|2020 | .................... | .................... | ..................... | .................... | .................... | .................... |
| V | 7496 hCoV-19/England/204690991/2020\|EPI_ISL_686592\|2020 | .................... | .................... | ..................... | .................... | .................... | .................... |

| | | | | | | | |
|---|---|---|---|---|---|---|---|
| V | 7497 hCoV-19/England/204691107/2020\|EPI_ISL_686593\|2020 | .................... | .A.................. | .................... | .................... | .................... | ...A................ |
| V | 7498 hCoV-19/England/204691117/2020\|EPI_ISL_686594\|2020 | .................... | .A.................. | .................... | .................... | .................... | ...A................ |
| V | 7499 hCoV-19/England/204691279/2020\|EPI_ISL_686595\|2020 | .................... | .................... | .................... | .................... | .................... | .................... |
| V | 7500 hCoV-19/England/204691372/2020\|EPI_ISL_686596\|2020 | .................... | .................... | .................... | .................... | .................... | .................... |
| V | 7501 hCoV-19/England/204691396/2020\|EPI_ISL_686598\|2020 | .................... | .................... | .................... | .................... | .................... | .................... |
| V | 7502 hCoV-19/England/204691655/2020\|EPI_ISL_686599\|2020 | .................... | .................... | .................... | .................... | .................... | .................... |
| V | 7503 hCoV-19/England/204821548_2/2020\|EPI_ISL_686611\|20 | .................... | .................... | .................... | .................... | .................... | .................... |
| V | 7504 hCoV-19/Japan/PG-7510/2020\|EPI_ISL_686612\|2020-07 | .................... | .................... | .................... | .................... | .................... | .................... |
| V | 7505 hCoV-19/Japan/PG-7511/2020\|EPI_ISL_686613\|2020-07 | .................... | .................... | .................... | .................... | .................... | .................... |
| V | 7506 hCoV-19/Japan/PG-7512/2020\|EPI_ISL_686614\|2020-08 | .................... | .................... | .................... | .................... | .................... | .................... |
| V | 7507 hCoV-19/Japan/PG-7513/2020\|EPI_ISL_686615\|2020-08 | .................... | .................... | .................... | .................... | .................... | .................... |
| V | 7508 hCoV-19/Japan/PG-7514/2020\|EPI_ISL_686616\|2020-08 | .................... | .................... | .................... | .................... | .................... | .................... |
| V | 7509 hCoV-19/Japan/PG-7515/2020\|EPI_ISL_686617\|2020-08 | .................... | .................... | .................... | .................... | .................... | .................... |
| V | 7510 hCoV-19/Japan/PG-7516/2020\|EPI_ISL_686618\|2020-08 | .................... | .................... | .................... | .................... | .................... | .................... |
| V | 7511 hCoV-19/Japan/PG-7517/2020\|EPI_ISL_686619\|2020-08 | .................... | .................... | .................... | .................... | .................... | .................... |
| V | 7512 hCoV-19/Japan/PG-7518/2020\|EPI_ISL_686620\|2020-08 | .................... | .................... | .................... | .................... | .................... | .................... |
| V | 7513 hCoV-19/Japan/PG-7519/2020\|EPI_ISL_686621\|2020-08 | .................... | .................... | .................... | .................... | .................... | .................... |
| V | 7514 hCoV-19/Japan/PG-7520/2020\|EPI_ISL_686622\|2020-08 | .................... | .................... | .................... | .................... | .................... | .................... |
| V | 7515 hCoV-19/Japan/PG-7521/2020\|EPI_ISL_686623\|2020-08 | .................... | .................... | .................... | .................... | .................... | .................... |
| V | 7516 hCoV-19/Japan/PG-7522/2020\|EPI_ISL_686624\|2020-08 | .................... | .................... | .................... | .................... | .................... | .................... |
| V | 7517 hCoV-19/Japan/PG-7523/2020\|EPI_ISL_686625\|2020-08 | .................... | .................... | .................... | .................... | .................... | .................... |
| V | 7518 hCoV-19/Japan/PG-7524/2020\|EPI_ISL_686626\|2020-08 | .................... | .................... | .................... | .................... | .................... | .................... |
| V | 7519 hCoV-19/Japan/PG-7525/2020\|EPI_ISL_686627\|2020-08 | .................... | .................... | .................... | .................... | .................... | .................... |
| V | 7520 hCoV-19/Japan/PG-7526/2020\|EPI_ISL_686628\|2020-08 | .................... | .................... | .................... | .................... | .................... | .................... |
| V | 7521 hCoV-19/Japan/PG-7527/2020\|EPI_ISL_686629\|2020-08 | .................... | .................... | .................... | .................... | .................... | .................... |
| V | 7522 hCoV-19/Japan/PG-7528/2020\|EPI_ISL_686630\|2020-08 | .................... | .................... | .................... | .................... | .................... | .................... |
| V | 7523 hCoV-19/Japan/PG-7529/2020\|EPI_ISL_686631\|2020-08 | .................... | .................... | .................... | .................... | .................... | .................... |
| V | 7524 hCoV-19/Japan/PG-7530/2020\|EPI_ISL_686632\|2020-08 | .................... | .................... | .................... | .................... | .................... | .................... |
| V | 7525 hCoV-19/Japan/PG-7531/2020\|EPI_ISL_686633\|2020-08 | .................... | .................... | .................... | .................... | .................... | .................... |
| V | 7526 hCoV-19/Japan/PG-7532/2020\|EPI_ISL_686634\|2020-08 | .................... | .................... | .................... | .................... | .................... | .................... |
| V | 7527 hCoV-19/Japan/PG-7533/2020\|EPI_ISL_686635\|2020-08 | .................... | .................... | .................... | .................... | .................... | .................... |
| V | 7528 hCoV-19/Japan/PG-7534/2020\|EPI_ISL_686636\|2020-08 | .................... | .................... | .................... | .................... | .................... | .................... |
| V | 7529 hCoV-19/Japan/PG-7535/2020\|EPI_ISL_686637\|2020-08 | .................... | .................... | .................... | .................... | .................... | .................... |
| V | 7530 hCoV-19/Japan/PG-7536/2020\|EPI_ISL_686638\|2020-08 | .................... | .................... | .................... | .................... | .................... | .................... |
| V | 7531 hCoV-19/Japan/PG-6834/2020\|EPI_ISL_687942\|2020-08 | .................... | .................... | .................... | .................... | .................... | .................... |
| V | 7532 hCoV-19/Japan/PG-6835/2020\|EPI_ISL_687943\|2020-08 | .................... | .................... | .................... | .................... | .................... | .................... |
| V | 7533 hCoV-19/Japan/PG-6836/2020\|EPI_ISL_687944\|2020-09 | .................... | .................... | .................... | .................... | .................... | .................... |
| V | 7534 hCoV-19/Japan/PG-6837/2020\|EPI_ISL_687945\|2020-09 | .................... | .................... | .................... | .................... | .................... | .................... |
| V | 7535 hCoV-19/Japan/PG-6838/2020\|EPI_ISL_687946\|2020-09 | .................... | .................... | .................... | .................... | .................... | .................... |
| V | 7536 hCoV-19/Japan/PG-6839/2020\|EPI_ISL_687947\|2020-09 | .................... | .................... | .................... | .................... | .................... | .................... |
| V | 7537 hCoV-19/Japan/PG-6840/2020\|EPI_ISL_687948\|2020-09 | .................... | .................... | .................... | .................... | .................... | .................... |
| V | 7538 hCoV-19/Japan/PG-6841/2020\|EPI_ISL_687949\|2020-09 | .................... | .................... | .................... | .................... | .................... | .................... |
| V | 7539 hCoV-19/Japan/PG-6842/2020\|EPI_ISL_687950\|2020-09 | .................... | .................... | .................... | .................... | .................... | .................... |
| V | 7540 hCoV-19/Japan/PG-6843/2020\|EPI_ISL_687951\|2020-09 | .................... | .................... | .................... | .................... | .................... | .................... |
| V | 7541 hCoV-19/Japan/PG-6844/2020\|EPI_ISL_687952\|2020-09 | .................... | .................... | .................... | .................... | .................... | .................... |
| V | 7542 hCoV-19/Japan/PG-6845/2020\|EPI_ISL_687953\|2020-09 | .................... | .................... | .................... | .................... | .................... | .................... |
| V | 7543 hCoV-19/Japan/PG-6846/2020\|EPI_ISL_687954\|2020-09 | .................... | .................... | .................... | .................... | .................... | .................... |
| V | 7544 hCoV-19/Japan/PG-6847/2020\|EPI_ISL_687955\|2020-09 | .................... | .................... | .................... | .................... | .................... | .................... |
| V | 7545 hCoV-19/Japan/PG-6848/2020\|EPI_ISL_687956\|2020-09 | .................... | .................... | .................... | .................... | .................... | .................... |
| V | 7546 hCoV-19/Japan/PG-6849/2020\|EPI_ISL_687957\|2020-09 | .................... | .................... | .................... | .................... | .................... | .................... |
| V | 7547 hCoV-19/Japan/PG-6850/2020\|EPI_ISL_687958\|2020-09 | .................... | .................... | .................... | .................... | .................... | .................... |
| V | 7548 hCoV-19/Japan/PG-6851/2020\|EPI_ISL_687959\|2020-09 | .................... | .................... | .................... | .................... | .................... | .................... |
| V | 7549 hCoV-19/Japan/PG-6852/2020\|EPI_ISL_687960\|2020-09 | .................... | .................... | .................... | .................... | .................... | .................... |
| V | 7550 hCoV-19/Japan/PG-6853/2020\|EPI_ISL_687961\|2020-09 | .................... | .................... | .................... | .................... | .................... | .................... |
| V | 7551 hCoV-19/Japan/PG-6854/2020\|EPI_ISL_687962\|2020-09 | .................... | .................... | .................... | .................... | .................... | .................... |
| V | 7552 hCoV-19/Japan/PG-6855/2020\|EPI_ISL_687963\|2020-09 | .................... | .................... | .................... | .................... | .................... | .................... |
| V | 7553 hCoV-19/Japan/PG-6856/2020\|EPI_ISL_687964\|2020-09 | .................... | .................... | .................... | .................... | .................... | .................... |
| V | 7554 hCoV-19/Japan/PG-6857/2020\|EPI_ISL_687965\|2020-09 | .................... | .................... | .................... | .................... | .................... | .................... |
| V | 7555 hCoV-19/Japan/PG-6858/2020\|EPI_ISL_687966\|2020-09 | .................... | .................... | .................... | .................... | .................... | .................... |
| V | 7556 hCoV-19/Japan/PG-6859/2020\|EPI_ISL_687967\|2020-09 | .................... | .................... | .................... | .................... | .................... | .................... |
| V | 7557 hCoV-19/Japan/PG-6860/2020\|EPI_ISL_687968\|2020-09 | .................... | .................... | .................... | .................... | .................... | .................... |
| V | 7558 hCoV-19/Japan/PG-6861/2020\|EPI_ISL_687969\|2020-09 | .................... | .................... | .................... | .................... | .................... | .................... |
| V | 7559 hCoV-19/Japan/PG-2975/2020\|EPI_ISL_691100\|2020-04 | .................... | .................... | .................... | .................... | .................... | .................... |
| V | 7560 hCoV-19/Japan/PG-2976/2020\|EPI_ISL_691101\|2020-04 | .................... | .................... | .................... | .................... | .................... | .................... |
| V | 7561 hCoV-19/Japan/PG-2977/2020\|EPI_ISL_691102\|2020-04 | .................... | .................... | .................... | .................... | .................... | .................... |
| V | 7562 hCoV-19/Japan/PG-2979/2020\|EPI_ISL_691103\|2020-04 | .................... | .................... | .................... | .................... | .................... | .................... |
| V | 7563 hCoV-19/Japan/PG-2980/2020\|EPI_ISL_691104\|2020-04 | .................... | .................... | .................... | .................... | .................... | .................... |
| V | 7564 hCoV-19/Japan/PG-2981/2020\|EPI_ISL_691105\|2020-05 | .................... | .................... | .................... | .................... | .................... | .................... |
| V | 7565 hCoV-19/Japan/PG-2982/2020\|EPI_ISL_691106\|2020-05 | .................... | .................... | .................... | .................... | .................... | .................... |
| V | 7566 hCoV-19/Japan/PG-2983/2020\|EPI_ISL_691107\|2020-05 | .................... | .................... | .................... | .................... | .................... | .................... |
| V | 7567 hCoV-19/Japan/PG-2984/2020\|EPI_ISL_691108\|2020-05 | .................... | .................... | .................... | .................... | .................... | .................... |
| V | 7569 hCoV-19/Japan/PG-2986/2020\|EPI_ISL_691110\|2020-04 | .................... | .................... | .................... | .................... | .................... | .................... |
| V | 7570 hCoV-19/Japan/PG-2987/2020\|EPI_ISL_691111\|2020-04 | .................... | .................... | .................... | .................... | .................... | .................... |
| V | 7571 hCoV-19/Japan/PG-2988/2020\|EPI_ISL_691112\|2020-04 | .................... | .................... | .................... | .................... | .................... | .................... |
| V | 7572 hCoV-19/Japan/PG-2989/2020\|EPI_ISL_691113\|2020-04 | .................... | .................... | .................... | .................... | .................... | .................... |
| V | 7573 hCoV-19/Japan/PG-2994/2020\|EPI_ISL_691118\|2020-04 | .................... | .................... | .................... | .................... | .................... | .................... |
| V | 7574 hCoV-19/Japan/PG-2995/2020\|EPI_ISL_691119\|2020-04 | .................... | .................... | .................... | .................... | .................... | .................... |
| V | 7575 hCoV-19/Japan/PG-3064/2020\|EPI_ISL_691187\|2020-04 | .................... | .................... | .................... | .................... | .................... | .................... |
| V | 7576 hCoV-19/Japan/PG-3065/2020\|EPI_ISL_691188\|2020-04 | .................... | .................... | .................... | .................... | .................... | .................... |
| V | 7577 hCoV-19/Japan/PG-3066/2020\|EPI_ISL_691189\|2020-04 | .................... | .................... | .................... | .................... | .................... | .................... |

V 7578 hCoV-19/Japan/PG-3067/2020|EPI_ISL_691190|2020-04 ···················· ···················· ····················· ···················· ···················· ····················
V 7579 hCoV-19/Japan/PG-3068/2020|EPI_ISL_691191|2020-04 ···················· ···················· ····················· ···················· ···················· ····················
V 7580 hCoV-19/Japan/PG-3069/2020|EPI_ISL_691192|2020-04 ···················· ···················· ····················· ···················· ···················· ····················
V 7581 hCoV-19/Japan/PG-3070/2020|EPI_ISL_691193|2020-04 ···················· ···················· ····················· ···················· ···················· ····················
V 7582 hCoV-19/Japan/PG-3071/2020|EPI_ISL_691194|2020-04 ···················· ···················· ····················· ···················· ···················· ····················
V 7583 hCoV-19/Japan/PG-3072/2020|EPI_ISL_691195|2020-04 ···················· ···················· ····················· ···················· ···················· ····················
V 7584 hCoV-19/Japan/PG-3073/2020|EPI_ISL_691196|2020-04 ···················· ···················· ····················· ···················· ···················· ····················
V 7585 hCoV-19/Japan/PG-3074/2020|EPI_ISL_691197|2020-04 ···················· ···················· ····················· ···················· ···················· ····················
V 7586 hCoV-19/Japan/PG-3075/2020|EPI_ISL_691198|2020-04 ···················· ···················· ····················· ···················· ···················· ····················
V 7587 hCoV-19/Japan/PG-3076/2020|EPI_ISL_691199|2020-04 ···················· ···················· ····················· ···················· ···················· ····················
V 7588 hCoV-19/Japan/PG-4161/2020|EPI_ISL_692400|2020-07 ···················· ···················· ····················· ···················· ···················· ····················
V 7589 hCoV-19/Japan/PG-4162/2020|EPI_ISL_692401|2020-07 ···················· ···················· ····················· ···················· ···················· ····················
V 7590 hCoV-19/Japan/PG-4163/2020|EPI_ISL_692402|2020-07 ···················· ···················· ····················· ···················· ···················· ····················
V 7591 hCoV-19/Japan/PG-4164/2020|EPI_ISL_692403|2020-07 ···················· ···················· ····················· ···················· ···················· ····················
V 7592 hCoV-19/Japan/PG-4165/2020|EPI_ISL_692404|2020-07 ···················· ···················· ····················· ···················· ···················· ····················
V 7593 hCoV-19/Japan/PG-4166/2020|EPI_ISL_692405|2020-07 ···················· ···················· ····················· ···················· ···················· ····················
V 7594 hCoV-19/Japan/PG-4167/2020|EPI_ISL_692406|2020-07 ···················· ···················· ····················· ···················· ···················· ····················
V 7595 hCoV-19/Japan/PG-4168/2020|EPI_ISL_692407|2020-07 ···················· ···················· ····················· ···················· ···················· ····················
V 7596 hCoV-19/Japan/PG-4169/2020|EPI_ISL_692408|2020-07 ···················· ···················· ····················· ···················· ···················· ····················
V 7597 hCoV-19/Japan/PG-4170/2020|EPI_ISL_692409|2020-07 ···················· ···················· ····················· ···················· ···················· ····················
V 7598 hCoV-19/Japan/PG-4171/2020|EPI_ISL_692410|2020-07 ···················· ···················· ····················· ···················· ···················· ····················
V 7599 hCoV-19/Japan/PG-4172/2020|EPI_ISL_692411|2020-07 ···················· ···················· ····················· ···················· ···················· ····················
V 7600 hCoV-19/Japan/PG-4173/2020|EPI_ISL_692412|2020-07 ···················· ···················· ····················· ···················· ···················· ····················
V 7601 hCoV-19/Japan/PG-4174/2020|EPI_ISL_692413|2020-07 ···················· ···················· ····················· ···················· ···················· ····················
V 7602 hCoV-19/Japan/PG-4175/2020|EPI_ISL_692414|2020-07 ···················· ···················· ····················· ···················· ···················· ····················
V 7603 hCoV-19/Japan/PG-4176/2020|EPI_ISL_692415|2020-07 ···················· ···················· ····················· ···················· ···················· ····················
V 7604 hCoV-19/Japan/PG-4177/2020|EPI_ISL_692416|2020-07 ···················· ···················· ····················· ···················· ···················· ····················
V 7605 hCoV-19/Japan/PG-4178/2020|EPI_ISL_692417|2020-07 ···················· ···················· ····················· ···················· ···················· ····················
V 7606 hCoV-19/Japan/PG-4179/2020|EPI_ISL_692418|2020-07 ···················· ···················· ····················· ···················· ···················· ····················
V 7607 hCoV-19/Japan/PG-4180/2020|EPI_ISL_692419|2020-07 ···················· ···················· ····················· ···················· ···················· ····················
V 7608 hCoV-19/Japan/PG-4181/2020|EPI_ISL_692420|2020-07 ···················· ···················· ····················· ···················· ···················· ····················
V 7609 hCoV-19/Japan/PG-4182/2020|EPI_ISL_692421|2020-07 ···················· ···················· ····················· ···················· ···················· ····················
V 7610 hCoV-19/Japan/PG-4183/2020|EPI_ISL_692422|2020-07 ···················· ···················· ····················· ···················· ···················· ····················
V 7611 hCoV-19/Japan/PG-4184/2020|EPI_ISL_692423|2020-07 ···················· ···················· ····················· ···················· ···················· ····················
V 7612 hCoV-19/Japan/PG-4185/2020|EPI_ISL_692424|2020-07 ···················· ···················· ····················· ···················· ···················· ····················
V 7613 hCoV-19/Japan/PG-4186/2020|EPI_ISL_692425|2020-07 ···················· ···················· ····················· ···················· ···················· ····················
V 7614 hCoV-19/Japan/PG-4187/2020|EPI_ISL_692426|2020-07 ···················· ···················· ····················· ···················· ···················· ····················
V 7615 hCoV-19/Japan/PG-4188/2020|EPI_ISL_692427|2020-07 ···················· ···················· ····················· ···················· ···················· ····················
V 7616 hCoV-19/Japan/PG-4189/2020|EPI_ISL_692428|2020-07 ···················· ···················· ····················· ···················· ···················· ····················
V 7617 hCoV-19/Japan/PG-4190/2020|EPI_ISL_692429|2020-07 ···················· ···················· ····················· ···················· ···················· ····················
V 7618 hCoV-19/Japan/PG-4191/2020|EPI_ISL_692430|2020-07 ···················· ···················· ····················· ···················· ···················· ····················
V 7619 hCoV-19/Japan/PG-4192/2020|EPI_ISL_692431|2020-07 ···················· ···················· ····················· ···················· ···················· ····················
V 7620 hCoV-19/Japan/PG-4193/2020|EPI_ISL_692432|2020-07 ···················· ···················· ····················· ···················· ···················· ····················
V 7621 hCoV-19/Japan/PG-4194/2020|EPI_ISL_692433|2020-07 ···················· ···················· ····················· ···················· ···················· ····················
V 7622 hCoV-19/Japan/PG-4195/2020|EPI_ISL_692434|2020-07 ···················· ···················· ····················· ···················· ···················· ····················
V 7623 hCoV-19/Japan/PG-4196/2020|EPI_ISL_692435|2020-07 ···················· ···················· ····················· ···················· ···················· ····················
V 7624 hCoV-19/Japan/PG-4197/2020|EPI_ISL_692436|2020-07 ···················· ···················· ····················· ···················· ···················· ····················
V 7625 hCoV-19/Japan/PG-4198/2020|EPI_ISL_692437|2020-07 ···················· ···················· ····················· ···················· ···················· ····················
V 7626 hCoV-19/Japan/PG-4200/2020|EPI_ISL_692438|2020-07 ···················· ···················· ····················· ···················· ···················· ····················
V 7627 hCoV-19/Japan/PG-4201/2020|EPI_ISL_692439|2020-07 ···················· ···················· ····················· ···················· ···················· ····················
V 7628 hCoV-19/Japan/PG-4202/2020|EPI_ISL_692440|2020-07 ···················· ···················· ····················· ···················· ···················· ····················
V 7629 hCoV-19/Japan/PG-4203/2020|EPI_ISL_692441|2020-07 ···················· ···················· ····················· ···················· ···················· ····················
V 7630 hCoV-19/Japan/PG-4204/2020|EPI_ISL_692442|2020-07 ···················· ···················· ····················· ···················· ···················· ····················
V 7631 hCoV-19/Japan/PG-4205/2020|EPI_ISL_692443|2020-07 ···················· ···················· ····················· ···················· ···················· ····················
V 7632 hCoV-19/Japan/PG-4206/2020|EPI_ISL_692444|2020-07 ···················· ···················· ····················· ···················· ···················· ····················
V 7633 hCoV-19/Japan/PG-4211/2020|EPI_ISL_692449|2020-07 ···················· ···················· ····················· ···················· ···················· ····················
V 7634 hCoV-19/USA/DE-DHSS-F970834/2020|EPI_ISL_693728|20 ···················· ···················· ····················· ···················· ···················· ····················
V 7635 hCoV-19/USA/DE-DHSS-F980493/2020|EPI_ISL_693729|20 ···················· ·········A·········· ····················· ···················· ···················· ····················
V 7636 hCoV-19/USA/DE-DHSS-F956496/2020|EPI_ISL_693730|20 ···················· ···················· ····················· ···················· ···················· ····················
V 7637 hCoV-19/USA/DE-DHSS-F956497/2020|EPI_ISL_693731|20 ···················· ···················· ····················· ···················· ···················· ····················
V 7638 hCoV-19/USA/DE-DHSS-F956498/2020|EPI_ISL_693732|20 ···················· ···················· ····················· ···················· ···················· ····················
V 7639 hCoV-19/USA/DE-DHSS-F956499/2020|EPI_ISL_693733|20 ···················· ···················· ····················· ···················· ···················· ····················
V 7640 hCoV-19/USA/DE-DHSS-F956500/2020|EPI_ISL_693734|20 ···················· ···················· ····················· ···················· ···················· ····················
V 7641 hCoV-19/USA/DE-DHSS-F956502/2020|EPI_ISL_693735|20 ···················· ···················· ····················· ···················· ···················· ····················
V 7642 hCoV-19/USA/DE-DHSS-F956503/2020|EPI_ISL_693736|20 ···················· ···················· ····················· ···················· ···················· ····················
V 7643 hCoV-19/USA/DE-DHSS-F956507/2020|EPI_ISL_693737|20 ···················· ···················· ····················· ···················· ···················· ····················
V 7644 hCoV-19/USA/DE-DHSS-F956508/2020|EPI_ISL_693738|20 ···················· ···················· ····················· ···················· ···················· ····················
V 7645 hCoV-19/USA/DE-DHSS-F956509/2020|EPI_ISL_693739|20 ···················· ···················· ····················· ···················· ···················· ····················
V 7646 hCoV-19/USA/DE-DHSS-F956511/2020|EPI_ISL_693740|20 ···················· ···················· ····················· ···················· ···················· ····················
V 7647 hCoV-19/USA/DE-DHSS-F957085/2020|EPI_ISL_693741|20 ···················· ···················· ····················· ···················· ···················· ····················
V 7648 hCoV-19/USA/DE-DHSS-F975901/2020|EPI_ISL_693742|20 ···················· ···················· ····················· ···················· ···················· ····················
V 7649 hCoV-19/USA/DE-DHSS-F981541/2020|EPI_ISL_693743|20 ···················· ···················· ····················· ···················· ···················· ····················
V 7650 hCoV-19/USA/DE-DHSS-F995183/2020|EPI_ISL_693744|20 ···················· ···················· ····················· ···················· ···················· ····················
V 7651 hCoV-19/USA/DE-DHSS-F1003822/2020|EPI_ISL_693745|2 ···················· ·A·················· ····················· ···················· ···················· ····················
V 7652 hCoV-19/USA/DE-DHSS-F1007797/2020|EPI_ISL_693746|2 ···················· ···················· ····················· ···················· ···················· ····················
V 7653 hCoV-19/USA/DE-DHSS-F956390/2020|EPI_ISL_693747|20 ···················· ···················· ····················· ···················· ···················· ····················
V 7654 hCoV-19/USA/DE-DHSS-F970674/2020|EPI_ISL_693748|20 ···················· ···················· ····················· ···················· ···················· ····················
V 7655 hCoV-19/USA/DE-DHSS-F970809/2020|EPI_ISL_693749|20 ···················· ···················· ····················· ···················· ···················· ····················
V 7656 hCoV-19/USA/DE-DHSS-F970817/2020|EPI_ISL_693750|20 ···················· ···················· ····················· ···················· ···················· ····················
V 7657 hCoV-19/USA/DE-DHSS-F995385/2020|EPI_ISL_693751|20 ···················· ·A·················· ····················· ···················· ···················· ····················

| | | | | | | | |
|---|---|---|---|---|---|---|---|
| V | 7658 hCoV-19/USA/DE-DHSS-F1009547/2020\|EPI_ISL_693752\|2 | ···················· | ···················· | ···················· | ···················· | ···················· | ···················· |
| V | 7659 hCoV-19/USA/DE-DHSS-F956506/2020\|EPI_ISL_693753\|20 | ···················· | ···················· | ···················· | ···················· | ···················· | ···················· |
| V | 7660 hCoV-19/USA/DE-DHSS-F970937/2020\|EPI_ISL_693754\|20 | ···················· | ···················· | ···················· | ···················· | ···················· | ···················· |
| V | 7661 hCoV-19/USA/DE-DHSS-F970939/2020\|EPI_ISL_693755\|20 | ···················· | ···················· | ···················· | ···················· | ···················· | ···················· |
| V | 7662 hCoV-19/USA/DE-DHSS-F980184/2020\|EPI_ISL_693756\|20 | ···················· | ···················· | ···················· | ···················· | ···················· | ···················· |
| V | 7663 hCoV-19/USA/DE-DHSS-F995589/2020\|EPI_ISL_693757\|20 | ···················· | ···················· | ···················· | ···················· | ···················· | ···················· |
| V | 7664 hCoV-19/France/BRE-IPP9944/2020\|EPI_ISL_693758\|202 | ···················· | ···················· | ···················· | ···················· | ···················· | ···················· |
| V | 7665 hCoV-19/France/BRE-IPP9945/2020\|EPI_ISL_693759\|202 | ···················· | ···················· | ···················· | ···················· | ···················· | ···················· |
| V | 7666 hCoV-19/France/BRE-IPP9946/2020\|EPI_ISL_693760\|202 | ···················· | ···················· | ···················· | ···················· | ···················· | ···················· |
| V | 7667 hCoV-19/France/BRE-IPP9949/2020\|EPI_ISL_693761\|202 | ···················· | ···················· | ···················· | ···················· | ···················· | ···················· |
| V | 7668 hCoV-19/France/BRE-IPP9954/2020\|EPI_ISL_693762\|202 | ···················· | ···················· | ···················· | ···················· | ···················· | ···················· |
| V | 7669 hCoV-19/France/BRE-IPP9957/2020\|EPI_ISL_693764\|202 | ···················· | ···················· | ···················· | ···················· | ···················· | ···················· |
| V | 7670 hCoV-19/France/BRE-IPP9961/2020\|EPI_ISL_693765\|202 | ···················· | ···················· | ···················· | ···················· | ···················· | ···················· |
| V | 7671 hCoV-19/France/CVL-IPP9875/2020\|EPI_ISL_693766\|202 | ···················· | ···················· | ···················· | ···················· | ···················· | ···················· |
| V | 7672 hCoV-19/France/GES-IPP9562/2020\|EPI_ISL_693767\|202 | ···················· | ···················· | ···················· | ···················· | ···················· | ···················· |
| V | 7673 hCoV-19/France/IDF-IPP9592/2020\|EPI_ISL_693768\|202 | ···················· | ···················· | ···················· | ···················· | ···················· | ···················· |
| V | 7674 hCoV-19/France/IDF-IPP9602/2020\|EPI_ISL_693769\|202 | ···················· | ···················· | ···················· | ···················· | ···················· | ···················· |
| V | 7675 hCoV-19/France/IDF-IPP9725/2020\|EPI_ISL_693770\|202 | ···················· | ···················· | ···················· | ···················· | ···················· | ···················· |
| V | 7676 hCoV-19/France/IDF-IPP9608/2020\|EPI_ISL_693771\|202 | ···················· | ···················· | ···················· | ···················· | ···················· | ···················· |
| V | 7677 hCoV-19/France/HDF-IPP9631/2020\|EPI_ISL_693772\|202 | ···················· | ···················· | ···················· | ···················· | ···················· | ···················· |
| V | 7678 hCoV-19/France/HDF-IPP9591/2020\|EPI_ISL_693773\|202 | ···················· | ···················· | ···················· | ···················· | ···················· | ···················· |
| V | 7679 hCoV-19/France/HDF-IPP9877/2020\|EPI_ISL_693774\|202 | ···················· | ···················· | ···················· | ···················· | ···················· | ···················· |
| V | 7680 hCoV-19/Switzerland/VS-ETHZ-350000/2020\|EPI_ISL_69 | ···················· | ···················· | ···················· | ···················· | ···················· | ···················· |
| V | 7681 hCoV-19/Japan/PG-1604/2020\|EPI_ISL_685210\|2020-04 | ···················· | ···················· | ···················· | ···················· | ···················· | ···················· |
| V | 7682 hCoV-19/Japan/PG-1605/2020\|EPI_ISL_685211\|2020-04 | ···················· | ···················· | ···················· | ···················· | ···················· | ···················· |
| V | 7683 hCoV-19/Japan/PG-1606/2020\|EPI_ISL_685212\|2020-04 | ···················· | ···················· | ···················· | ···················· | ···················· | ···················· |
| V | 7684 hCoV-19/Japan/PG-1607/2020\|EPI_ISL_685213\|2020-04 | ···················· | ···················· | ···················· | ···················· | ···················· | ···················· |
| V | 7685 hCoV-19/Japan/PG-1608/2020\|EPI_ISL_685214\|2020-04 | ···················· | ···················· | ···················· | ···················· | ···················· | ···················· |
| V | 7686 hCoV-19/Japan/PG-1609/2020\|EPI_ISL_685215\|2020-04 | ···················· | ···················· | ···················· | ···················· | ···················· | ···················· |
| V | 7687 hCoV-19/Japan/PG-1610/2020\|EPI_ISL_685216\|2020-04 | ···················· | ···················· | ···················· | ···················· | ···················· | ···················· |
| V | 7688 hCoV-19/Japan/PG-1611/2020\|EPI_ISL_685217\|2020-04 | ···················· | ···················· | ···················· | ···················· | ···················· | ···················· |
| V | 7689 hCoV-19/Japan/PG-1612/2020\|EPI_ISL_685218\|2020-04 | ···················· | ···················· | ···················· | ···················· | ···················· | ···················· |
| V | 7690 hCoV-19/Japan/PG-1613/2020\|EPI_ISL_685219\|2020-04 | ···················· | ···················· | ···················· | ···················· | ···················· | ···················· |
| V | 7691 hCoV-19/Japan/PG-1614/2020\|EPI_ISL_685220\|2020-03 | ···················· | ···················· | ···················· | ···················· | ···················· | ···················· |
| V | 7692 hCoV-19/Japan/PG-1615/2020\|EPI_ISL_685221\|2020-04 | ···················· | ···················· | ···················· | ···················· | ···················· | ···················· |
| V | 7693 hCoV-19/Japan/PG-1616/2020\|EPI_ISL_685222\|2020-04 | ···················· | ···················· | ···················· | ···················· | ···················· | ···················· |
| V | 7694 hCoV-19/Japan/PG-1617/2020\|EPI_ISL_685223\|2020-04 | ···················· | ···················· | ···················· | ···················· | ···················· | ···················· |
| V | 7695 hCoV-19/Japan/PG-1618/2020\|EPI_ISL_685224\|2020-04 | ···················· | ···················· | ···················· | ···················· | ···················· | ···················· |
| V | 7696 hCoV-19/Japan/PG-1619/2020\|EPI_ISL_685225\|2020-04 | ···················· | ···················· | ···················· | ···················· | ···················· | ···················· |
| V | 7697 hCoV-19/Japan/PG-1620/2020\|EPI_ISL_685226\|2020-04 | ···················· | ···················· | ···················· | ···················· | ···················· | ···················· |
| V | 7698 hCoV-19/Japan/PG-1621/2020\|EPI_ISL_685227\|2020-04 | ···················· | ···················· | ···················· | ···················· | ···················· | ···················· |
| V | 7699 hCoV-19/Japan/PG-1622/2020\|EPI_ISL_685228\|2020-04 | ···················· | ···················· | ···················· | ···················· | ···················· | ···················· |
| V | 7700 hCoV-19/Japan/PG-1623/2020\|EPI_ISL_685229\|2020-04 | ···················· | ···················· | ···················· | ···················· | ···················· | ···················· |
| V | 7701 hCoV-19/Japan/PG-1624/2020\|EPI_ISL_685230\|2020-04 | ···················· | ···················· | ···················· | ···················· | ···················· | ···················· |
| V | 7702 hCoV-19/Japan/PG-1625/2020\|EPI_ISL_685231\|2020-04 | ···················· | ···················· | ···················· | ···················· | ···················· | ···················· |
| V | 7703 hCoV-19/Japan/PG-1626/2020\|EPI_ISL_685232\|2020-04 | ···················· | ···················· | ···················· | ···················· | ···················· | ···················· |
| V | 7704 hCoV-19/Japan/PG-1627/2020\|EPI_ISL_685233\|2020-04 | ···················· | ···················· | ···················· | ···················· | ···················· | ···················· |
| V | 7705 hCoV-19/Japan/PG-1628/2020\|EPI_ISL_685234\|2020-04 | ···················· | ···················· | ···················· | ···················· | ···················· | ···················· |
| V | 7706 hCoV-19/Japan/PG-1629/2020\|EPI_ISL_685235\|2020-04 | ···················· | ···················· | ···················· | ···················· | ···················· | ···················· |
| V | 7707 hCoV-19/Japan/PG-1630/2020\|EPI_ISL_685236\|2020-04 | ···················· | ···················· | ···················· | ···················· | ···················· | ···················· |
| V | 7708 hCoV-19/Japan/PG-1631/2020\|EPI_ISL_685237\|2020-04 | ···················· | ···················· | ···················· | ···················· | ···················· | ···················· |
| V | 7709 hCoV-19/Japan/PG-1632/2020\|EPI_ISL_685238\|2020-03 | ···················· | ···················· | ···················· | ···················· | ···················· | ···················· |
| V | 7710 hCoV-19/Japan/PG-1634/2020\|EPI_ISL_685240\|2020-03 | ···················· | ···················· | ···················· | ···················· | ···················· | ···················· |
| V | 7711 hCoV-19/Japan/PG-1635/2020\|EPI_ISL_685241\|2020-03 | ···················· | ···················· | ···················· | ···················· | ···················· | ···················· |
| V | 7712 hCoV-19/Japan/PG-1636/2020\|EPI_ISL_685242\|2020-03 | ···················· | ···················· | ···················· | ···················· | ···················· | ···················· |
| V | 7713 hCoV-19/Japan/PG-1637/2020\|EPI_ISL_685243\|2020-03 | ···················· | ···················· | ···················· | ···················· | ···················· | ···················· |
| V | 7714 hCoV-19/Japan/PG-1638/2020\|EPI_ISL_685244\|2020-04 | ···················· | ···················· | ···················· | ···················· | ···················· | ···················· |
| V | 7715 hCoV-19/Japan/PG-1639/2020\|EPI_ISL_685245\|2020-04 | ···················· | ···················· | ···················· | ···················· | ···················· | ···················· |
| V | 7716 hCoV-19/Japan/PG-1640/2020\|EPI_ISL_685246\|2020-04 | ···················· | ···················· | ···················· | ···················· | ···················· | ···················· |
| V | 7717 hCoV-19/Japan/PG-1641/2020\|EPI_ISL_685247\|2020-04 | ···················· | ···················· | ···················· | ···················· | ···················· | ···················· |
| V | 7718 hCoV-19/Japan/PG-1642/2020\|EPI_ISL_685248\|2020-04 | ···················· | ···················· | ···················· | ···················· | ···················· | ···················· |
| V | 7719 hCoV-19/Japan/PG-1643/2020\|EPI_ISL_685249\|2020-04 | ···················· | ···················· | ···················· | ···················· | ···················· | ···················· |
| V | 7720 hCoV-19/England/204040039/2020\|EPI_ISL_686540\|2020 | ···················· | ···················· | ···················· | ···················· | ···················· | ···················· |
| V | 7721 hCoV-19/England/204540373/2020\|EPI_ISL_686543\|2020 | ···················· | ···················· | ···················· | ···················· | ···················· | ···················· |
| V | 7722 hCoV-19/England/204561454/2020\|EPI_ISL_686544\|2020 | ···················· | ·A·················· | ···················· | ···················· | ···················· | ··A················· |
| V | 7723 hCoV-19/England/204580253/2020\|EPI_ISL_686547\|2020 | ···················· | ···················· | ···················· | ···················· | ···················· | ···················· |
| V | 7724 hCoV-19/England/204581975/2020\|EPI_ISL_686548\|2020 | ···················· | ···················· | ···················· | ···················· | ···················· | ···················· |
| V | 7725 hCoV-19/England/204590229/2020\|EPI_ISL_686550\|2020 | ···················· | ···················· | ···················· | ···················· | ···················· | ···················· |
| V | 7726 hCoV-19/England/204590294/2020\|EPI_ISL_686551\|2020 | ···················· | ···················· | ···················· | ···················· | ···················· | ···················· |
| V | 7727 hCoV-19/England/204590330/2020\|EPI_ISL_686552\|2020 | ···················· | ···················· | ···················· | ···················· | ···················· | ···················· |
| V | 7728 hCoV-19/England/204590422/2020\|EPI_ISL_686553\|2020 | ···················· | ···················· | ···················· | ···················· | ···················· | ···················· |
| V | 7729 hCoV-19/England/204590455/2020\|EPI_ISL_686554\|2020 | ···················· | ···················· | ···················· | ···················· | ···················· | ···················· |
| V | 7730 hCoV-19/England/204590467/2020\|EPI_ISL_686555\|2020 | ···················· | ···················· | ···················· | ···················· | ···················· | ···················· |
| V | 7731 hCoV-19/England/204590468/2020\|EPI_ISL_686556\|2020 | ···················· | ···················· | ···················· | ···················· | ···················· | ···················· |
| V | 7732 hCoV-19/England/204590470/2020\|EPI_ISL_686557\|2020 | ···················· | ···················· | ···················· | ···················· | ···················· | ···················· |
| V | 7733 hCoV-19/England/204591424/2020\|EPI_ISL_686558\|2020 | ···················· | ···················· | ···················· | ···················· | ···················· | ···················· |
| V | 7734 hCoV-19/England/204591431/2020\|EPI_ISL_686559\|2020 | ···················· | ···················· | ···················· | ···················· | ···················· | ···················· |
| V | 7735 hCoV-19/England/204620444/2020\|EPI_ISL_686560\|2020 | ···················· | ···················· | ···················· | ···················· | ·····T·············· | ···················· |
| V | 7736 hCoV-19/England/204621581/2020\|EPI_ISL_686561\|2020 | ···················· | ···················· | ···················· | ···················· | ···················· | ···················· |
| V | 7737 hCoV-19/England/204641361/2020\|EPI_ISL_686562\|2020 | ···················· | ···················· | ···················· | ···················· | ···················· | ···················· |

| | | | | | | | |
|---|---|---|---|---|---|---|---|
| V | 7738 hCoV-19/England/204660291/2020\|EPI_ISL_686563\|2020 | ···················· | ···················· | ···················· | ···················· | ·····T·············· | ···················· |
| V | 7739 hCoV-19/England/204660292/2020\|EPI_ISL_686564\|2020 | ···················· | ···················· | ···················· | ···················· | ·····T·············· | ···················· |
| V | 7740 hCoV-19/England/204660298/2020\|EPI_ISL_686565\|2020 | ···················· | ···················· | ···················· | ···················· | ·····T·············· | ···················· |
| V | 7741 hCoV-19/England/204660300/2020\|EPI_ISL_686566\|2020 | ···················· | ···················· | ···················· | ···················· | ·····T·············· | ···················· |
| V | 7742 hCoV-19/England/204660311/2020\|EPI_ISL_686567\|2020 | ···················· | ···················· | ···················· | ···················· | ·····T·············· | ···················· |
| V | 7743 hCoV-19/England/204660312/2020\|EPI_ISL_686568\|2020 | ···················· | ···················· | ···················· | ···················· | ·····T·············· | ···················· |
| V | 7744 hCoV-19/England/204660314/2020\|EPI_ISL_686570\|2020 | ···················· | ···················· | ···················· | ···················· | ·····T·············· | ···················· |
| V | 7745 hCoV-19/England/204660318/2020\|EPI_ISL_686571\|2020 | ···················· | ···················· | ···················· | ···················· | ·····T·············· | ···················· |
| V | 7746 hCoV-19/England/204660319/2020\|EPI_ISL_686572\|2020 | ···················· | ···················· | ···················· | ···················· | ·····T·············· | ···················· |
| V | 7747 hCoV-19/England/204660322/2020\|EPI_ISL_686573\|2020 | ···················· | ···················· | ···················· | ···················· | ·····T·············· | ···················· |
| V | 7748 hCoV-19/England/204660323/2020\|EPI_ISL_686574\|2020 | ···················· | ···················· | ···················· | ···················· | ·····T·············· | ···················· |
| V | 7749 hCoV-19/England/204661388/2020\|EPI_ISL_686575\|2020 | ···················· | ···················· | ···················· | ···················· | ·····T·············· | ···················· |
| V | 7750 hCoV-19/England/204661389/2020\|EPI_ISL_686576\|2020 | ···················· | ···················· | ···················· | ···················· | ·····T·············· | ···················· |
| V | 7751 hCoV-19/England/204661392/2020\|EPI_ISL_686577\|2020 | ···················· | ···················· | ···················· | ···················· | ·····T·············· | ···················· |
| V | 7752 hCoV-19/England/204661397/2020\|EPI_ISL_686578\|2020 | ···················· | ···················· | ···················· | ···················· | ·····T·············· | ···················· |
| V | 7753 hCoV-19/England/204661400/2020\|EPI_ISL_686579\|2020 | ···················· | ···················· | ···················· | ···················· | ·····T·············· | ···················· |
| V | 7754 hCoV-19/Japan/PG-6689/2020\|EPI_ISL_687800\|2020-07 | ···················· | ···················· | ···················· | ···················· | ···················· | ···················· |
| V | 7755 hCoV-19/Japan/PG-6690/2020\|EPI_ISL_687801\|2020-07 | ···················· | ···················· | ···················· | ···················· | ···················· | ···················· |
| V | 7756 hCoV-19/Japan/PG-6691/2020\|EPI_ISL_687802\|2020-07 | ···················· | ···················· | ···················· | ···················· | ···················· | ···················· |
| V | 7757 hCoV-19/Japan/PG-6692/2020\|EPI_ISL_687803\|2020-07 | ···················· | ···················· | ···················· | ···················· | ···················· | ···················· |
| V | 7758 hCoV-19/Japan/PG-6693/2020\|EPI_ISL_687804\|2020-07 | ···················· | ···················· | ···················· | ···················· | ···················· | ···················· |
| V | 7759 hCoV-19/Japan/PG-6694/2020\|EPI_ISL_687805\|2020-07 | ···················· | ···················· | ···················· | ···················· | ···················· | ···················· |
| V | 7760 hCoV-19/Japan/PG-6695/2020\|EPI_ISL_687806\|2020-07 | ···················· | ···················· | ···················· | ···················· | ···················· | ···················· |
| V | 7761 hCoV-19/Japan/PG-6696/2020\|EPI_ISL_687807\|2020-07 | ···················· | ···················· | ···················· | ···················· | ···················· | ···················· |
| V | 7762 hCoV-19/Japan/PG-6697/2020\|EPI_ISL_687808\|2020-07 | ···················· | ···················· | ···················· | ···················· | ···················· | ···················· |
| V | 7763 hCoV-19/Japan/PG-6698/2020\|EPI_ISL_687809\|2020-07 | ···················· | ···················· | ···················· | ···················· | ···················· | ···················· |
| V | 7764 hCoV-19/Japan/PG-6699/2020\|EPI_ISL_687810\|2020-07 | ···················· | ···················· | ···················· | ···················· | ···················· | ···················· |
| V | 7765 hCoV-19/Japan/PG-6700/2020\|EPI_ISL_687811\|2020-07 | ···················· | ···················· | ···················· | ···················· | ···················· | ···················· |
| V | 7766 hCoV-19/Japan/PG-6701/2020\|EPI_ISL_687812\|2020-07 | ···················· | ···················· | ···················· | ···················· | ···················· | ···················· |
| V | 7767 hCoV-19/Japan/PG-6702/2020\|EPI_ISL_687813\|2020-07 | ···················· | ···················· | ···················· | ···················· | ···················· | ···················· |
| V | 7768 hCoV-19/Japan/PG-6703/2020\|EPI_ISL_687814\|2020-07 | ···················· | ···················· | ···················· | ···················· | ···················· | ···················· |
| V | 7769 hCoV-19/Japan/PG-6704/2020\|EPI_ISL_687815\|2020-07 | ···················· | ···················· | ···················· | ···················· | ···················· | ···················· |
| V | 7770 hCoV-19/Japan/PG-6705/2020\|EPI_ISL_687816\|2020-07 | ···················· | ···················· | ···················· | ···················· | ···················· | ···················· |
| V | 7771 hCoV-19/Japan/PG-6706/2020\|EPI_ISL_687817\|2020-07 | ···················· | ···················· | ···················· | ···················· | ···················· | ···················· |
| V | 7772 hCoV-19/Japan/PG-6707/2020\|EPI_ISL_687818\|2020-07 | ···················· | ···················· | ···················· | ···················· | ···················· | ···················· |
| V | 7773 hCoV-19/Japan/PG-6708/2020\|EPI_ISL_687819\|2020-07 | ···················· | ···················· | ···················· | ···················· | ···················· | ···················· |
| V | 7774 hCoV-19/Japan/PG-6709/2020\|EPI_ISL_687820\|2020-07 | ···················· | ···················· | ···················· | ···················· | ···················· | ···················· |
| V | 7775 hCoV-19/Japan/PG-6760/2020\|EPI_ISL_687870\|2020-08 | ···················· | ···················· | ···················· | ···················· | ···················· | ···················· |
| V | 7776 hCoV-19/Japan/PG-6761/2020\|EPI_ISL_687871\|2020-08 | ···················· | ···················· | ···················· | ···················· | ···················· | ···················· |
| V | 7777 hCoV-19/Japan/PG-6762/2020\|EPI_ISL_687872\|2020-08 | ···················· | ···················· | ···················· | ···················· | ···················· | ···················· |
| V | 7778 hCoV-19/Japan/PG-6763/2020\|EPI_ISL_687873\|2020-08 | ···················· | ···················· | ···················· | ···················· | ···················· | ···················· |
| V | 7779 hCoV-19/Japan/PG-6764/2020\|EPI_ISL_687874\|2020-08 | ···················· | ···················· | ···················· | ···················· | ···················· | ···················· |
| V | 7780 hCoV-19/Japan/PG-6765/2020\|EPI_ISL_687875\|2020-08 | ···················· | ···················· | ···················· | ···················· | ···················· | ···················· |
| V | 7781 hCoV-19/Japan/PG-6766/2020\|EPI_ISL_687876\|2020-08 | ···················· | ···················· | ···················· | ···················· | ···················· | ···················· |
| V | 7782 hCoV-19/Japan/PG-6767/2020\|EPI_ISL_687877\|2020-08 | ···················· | ···················· | ···················· | ···················· | ···················· | ···················· |
| V | 7783 hCoV-19/Japan/PG-6768/2020\|EPI_ISL_687878\|2020-08 | ···················· | ···················· | ···················· | ···················· | ···················· | ···················· |
| V | 7784 hCoV-19/Japan/PG-6769/2020\|EPI_ISL_687879\|2020-08 | ···················· | ···················· | ···················· | ···················· | ···················· | ···················· |
| V | 7785 hCoV-19/Japan/PG-6770/2020\|EPI_ISL_687880\|2020-08 | ···················· | ···················· | ···················· | ···················· | ···················· | ···················· |
| V | 7786 hCoV-19/Japan/PG-6771/2020\|EPI_ISL_687881\|2020-08 | ···················· | ···················· | ···················· | ···················· | ···················· | ···················· |
| V | 7787 hCoV-19/Japan/PG-6772/2020\|EPI_ISL_687882\|2020-08 | ···················· | ···················· | ···················· | ···················· | ···················· | ···················· |
| V | 7788 hCoV-19/Japan/PG-6773/2020\|EPI_ISL_687883\|2020-08 | ···················· | ···················· | ···················· | ···················· | ···················· | ···················· |
| V | 7789 hCoV-19/Japan/PG-6774/2020\|EPI_ISL_687884\|2020-08 | ···················· | ···················· | ···················· | ···················· | ···················· | ···················· |
| V | 7790 hCoV-19/Japan/PG-6775/2020\|EPI_ISL_687885\|2020-08 | ···················· | ···················· | ···················· | ···················· | ···················· | ···················· |
| V | 7791 hCoV-19/Japan/PG-6776/2020\|EPI_ISL_687886\|2020-08 | ···················· | ···················· | ···················· | ···················· | ···················· | ···················· |
| V | 7792 hCoV-19/Japan/PG-6777/2020\|EPI_ISL_687887\|2020-08 | ···················· | ···················· | ···················· | ···················· | ···················· | ···················· |
| V | 7793 hCoV-19/Japan/PG-6778/2020\|EPI_ISL_687888\|2020-08 | ···················· | ···················· | ···················· | ···················· | ···················· | ···················· |
| V | 7794 hCoV-19/Japan/PG-6780/2020\|EPI_ISL_687889\|2020-08 | ···················· | ···················· | ···················· | ···················· | ···················· | ···················· |
| V | 7795 hCoV-19/Japan/PG-6781/2020\|EPI_ISL_687890\|2020-08 | ···················· | ···················· | ···················· | ···················· | ···················· | ···················· |
| V | 7796 hCoV-19/Japan/PG-6782/2020\|EPI_ISL_687891\|2020-08 | ···················· | ···················· | ···················· | ···················· | ···················· | ···················· |
| V | 7797 hCoV-19/Japan/PG-6783/2020\|EPI_ISL_687892\|2020-08 | ···················· | ···················· | ···················· | ···················· | ···················· | ···················· |
| V | 7798 hCoV-19/Japan/PG-6784/2020\|EPI_ISL_687893\|2020-08 | ···················· | ···················· | ···················· | ···················· | ···················· | ···················· |
| V | 7799 hCoV-19/Japan/PG-6785/2020\|EPI_ISL_687894\|2020-08 | ···················· | ···················· | ···················· | ···················· | ···················· | ···················· |
| V | 7800 hCoV-19/Japan/PG-6786/2020\|EPI_ISL_687895\|2020-08 | ···················· | ···················· | ···················· | ···················· | ···················· | ···················· |
| V | 7801 hCoV-19/Japan/PG-6787/2020\|EPI_ISL_687896\|2020-08 | ···················· | ···················· | ···················· | ···················· | ···················· | ···················· |
| V | 7802 hCoV-19/Japan/PG-6788/2020\|EPI_ISL_687897\|2020-08 | ···················· | ···················· | ···················· | ···················· | ···················· | ···················· |
| V | 7803 hCoV-19/Japan/PG-6789/2020\|EPI_ISL_687898\|2020-08 | ···················· | ···················· | ···················· | ···················· | ···················· | ···················· |
| V | 7804 hCoV-19/Japan/PG-6790/2020\|EPI_ISL_687899\|2020-08 | ···················· | ···················· | ···················· | ···················· | ···················· | ···················· |
| V | 7805 hCoV-19/Japan/PG-2874/2020\|EPI_ISL_691000\|2020-04 | ···················· | ···················· | ···················· | ···················· | ···················· | ···················· |
| V | 7806 hCoV-19/Japan/PG-2875/2020\|EPI_ISL_691001\|2020-04 | ···················· | ···················· | ···················· | ···················· | ···················· | ···················· |
| V | 7807 hCoV-19/Japan/PG-2876/2020\|EPI_ISL_691002\|2020-04 | ···················· | ···················· | ···················· | ···················· | ···················· | ···················· |
| V | 7808 hCoV-19/Japan/PG-2877/2020\|EPI_ISL_691003\|2020-04 | ···················· | ···················· | ···················· | ···················· | ···················· | ···················· |
| V | 7809 hCoV-19/Japan/PG-2878/2020\|EPI_ISL_691004\|2020-04 | ···················· | ···················· | ···················· | ···················· | ···················· | ···················· |
| V | 7810 hCoV-19/Japan/PG-2879/2020\|EPI_ISL_691005\|2020-04 | ···················· | ···················· | ···················· | ···················· | ···················· | ···················· |
| V | 7811 hCoV-19/Japan/PG-2880/2020\|EPI_ISL_691006\|2020-04 | ···················· | ···················· | ···················· | ···················· | ···················· | ···················· |
| V | 7812 hCoV-19/Japan/PG-2881/2020\|EPI_ISL_691007\|2020-04 | ···················· | ···················· | ···················· | ···················· | ···················· | ···················· |
| V | 7813 hCoV-19/Japan/PG-2882/2020\|EPI_ISL_691008\|2020-04 | ···················· | ···················· | ···················· | ···················· | ···················· | ···················· |
| V | 7814 hCoV-19/Japan/PG-2883/2020\|EPI_ISL_691009\|2020-04 | ···················· | ···················· | ···················· | ···················· | ···················· | ···················· |
| V | 7815 hCoV-19/Japan/PG-2884/2020\|EPI_ISL_691010\|2020-04 | ···················· | ···················· | ···················· | ···················· | ···················· | ···················· |
| V | 7817 hCoV-19/Japan/PG-2886/2020\|EPI_ISL_691012\|2020-05 | ···················· | ···················· | ···················· | ···················· | ···················· | ···················· |
| V | 7818 hCoV-19/Japan/PG-2887/2020\|EPI_ISL_691013\|2020-05 | ···················· | ···················· | ···················· | ···················· | ···················· | ···················· |

| | | | | | | | | |
|---|---|---|---|---|---|---|---|---|
| V | 7820 | hCoV-19/Japan/PG-2889/2020\|EPI_ISL_691015\|2020-03 | .................... | .................... | ..................... | .................... | .................... | .................... |
| V | 7821 | hCoV-19/Japan/PG-2890/2020\|EPI_ISL_691016\|2020-03 | .................... | .................... | ..................... | .................... | .................... | .................... |
| V | 7822 | hCoV-19/Japan/PG-2891/2020\|EPI_ISL_691017\|2020-03 | .................... | .................... | ..................... | .................... | .................... | .................... |
| V | 7823 | hCoV-19/Japan/PG-2892/2020\|EPI_ISL_691018\|2020-03 | .................... | .................... | ..................... | .................... | .................... | .................... |
| V | 7824 | hCoV-19/Japan/PG-2893/2020\|EPI_ISL_691019\|2020-03 | .................... | .................... | ..................... | .................... | .................... | .................... |
| V | 7825 | hCoV-19/Japan/PG-2894/2020\|EPI_ISL_691020\|2020-03 | .................... | .................... | ..................... | .................... | .................... | .................... |
| V | 7826 | hCoV-19/Japan/PG-2895/2020\|EPI_ISL_691021\|2020-03 | .................... | .................... | ..................... | .................... | .................... | .................... |
| V | 7827 | hCoV-19/Japan/PG-2896/2020\|EPI_ISL_691022\|2020-03 | .................... | .................... | ..................... | .................... | .................... | .................... |
| V | 7828 | hCoV-19/Japan/PG-2897/2020\|EPI_ISL_691023\|2020-03 | .................... | .................... | ..................... | .................... | .................... | .................... |
| V | 7829 | hCoV-19/Japan/PG-2898/2020\|EPI_ISL_691024\|2020-03 | .................... | .................... | ..................... | .................... | .................... | .................... |
| V | 7830 | hCoV-19/Japan/PG-2899/2020\|EPI_ISL_691025\|2020-03 | .................... | .................... | ..................... | .................... | .................... | .................... |
| V | 7831 | hCoV-19/Japan/PG-2900/2020\|EPI_ISL_691026\|2020-03 | .................... | .................... | ..................... | .................... | .................... | .................... |
| V | 7832 | hCoV-19/Japan/PG-2901/2020\|EPI_ISL_691027\|2020-03 | .................... | .................... | ..................... | .................... | .................... | .................... |
| V | 7833 | hCoV-19/Japan/PG-2902/2020\|EPI_ISL_691028\|2020-03 | .................... | .................... | ..................... | .................... | .................... | .................... |
| V | 7834 | hCoV-19/Japan/PG-2903/2020\|EPI_ISL_691029\|2020-03 | .................... | .................... | ..................... | .................... | .................... | .................... |
| V | 7835 | hCoV-19/Japan/PG-2904/2020\|EPI_ISL_691030\|2020-03 | .................... | .................... | ..................... | .................... | .................... | .................... |
| V | 7836 | hCoV-19/Japan/PG-2905/2020\|EPI_ISL_691031\|2020-03 | .................... | .................... | ..................... | .................... | .................... | .................... |
| V | 7837 | hCoV-19/Japan/PG-2906/2020\|EPI_ISL_691032\|2020-03 | .................... | .................... | ..................... | .................... | .................... | .................... |
| V | 7838 | hCoV-19/Japan/PG-2907/2020\|EPI_ISL_691033\|2020-04 | .................... | .................... | ..................... | .................... | .................... | .................... |
| V | 7839 | hCoV-19/Japan/PG-2908/2020\|EPI_ISL_691034\|2020-04 | .................... | .................... | ..................... | .................... | .................... | .................... |
| V | 7840 | hCoV-19/Japan/PG-2909/2020\|EPI_ISL_691035\|2020-04 | .................... | .................... | ..................... | .................... | .................... | .................... |
| V | 7841 | hCoV-19/Japan/PG-2910/2020\|EPI_ISL_691036\|2020-04 | .................... | .................... | ..................... | .................... | .................... | .................... |
| V | 7842 | hCoV-19/Japan/PG-2911/2020\|EPI_ISL_691037\|2020-04 | .................... | .................... | ..................... | .................... | .................... | .................... |
| V | 7843 | hCoV-19/Japan/PG-2913/2020\|EPI_ISL_691039\|2020-04 | .................... | .................... | ..................... | .................... | .................... | .................... |
| V | 7844 | hCoV-19/Japan/PG-2962/2020\|EPI_ISL_691088\|2020-04 | .................... | .................... | ..................... | .................... | .................... | .................... |
| V | 7845 | hCoV-19/Japan/PG-2963/2020\|EPI_ISL_691089\|2020-04 | .................... | .................... | ..................... | .................... | .................... | .................... |
| V | 7846 | hCoV-19/Japan/PG-2964/2020\|EPI_ISL_691090\|2020-04 | .................... | .................... | ..................... | .................... | .................... | .................... |
| V | 7847 | hCoV-19/Japan/PG-2965/2020\|EPI_ISL_691091\|2020-04 | .................... | .................... | ..................... | .................... | .................... | .................... |
| V | 7848 | hCoV-19/Japan/PG-2966/2020\|EPI_ISL_691092\|2020-04 | .................... | .................... | ..................... | .................... | .................... | .................... |
| V | 7849 | hCoV-19/Japan/PG-2967/2020\|EPI_ISL_691093\|2020-04 | .................... | .................... | ..................... | .................... | .................... | .................... |
| V | 7850 | hCoV-19/Japan/PG-2968/2020\|EPI_ISL_691094\|2020-04 | .................... | .................... | ..................... | .................... | .................... | .................... |
| V | 7851 | hCoV-19/Japan/PG-2970/2020\|EPI_ISL_691095\|2020-04 | .................... | .................... | ..................... | .................... | .................... | .................... |
| V | 7852 | hCoV-19/Japan/PG-2971/2020\|EPI_ISL_691096\|2020-04 | .................... | .................... | ..................... | .................... | .................... | .................... |
| V | 7853 | hCoV-19/Japan/PG-2972/2020\|EPI_ISL_691097\|2020-04 | .................... | .................... | ..................... | .................... | .................... | .................... |
| V | 7854 | hCoV-19/Japan/PG-2973/2020\|EPI_ISL_691098\|2020-04 | .................... | .................... | ..................... | .................... | .................... | .................... |
| V | 7855 | hCoV-19/Japan/PG-2974/2020\|EPI_ISL_691099\|2020-04 | .................... | .................... | ..................... | .................... | .................... | .................... |
| V | 7856 | hCoV-19/Japan/PG-2990/2020\|EPI_ISL_691114\|2020-04 | .................... | .................... | ..................... | .................... | .................... | .................... |
| V | 7857 | hCoV-19/Japan/PG-2991/2020\|EPI_ISL_691115\|2020-04 | .................... | .................... | ..................... | .................... | .................... | .................... |
| V | 7858 | hCoV-19/Japan/PG-2992/2020\|EPI_ISL_691116\|2020-04 | .................... | .................... | ..................... | .................... | .................... | .................... |
| V | 7859 | hCoV-19/Japan/PG-2993/2020\|EPI_ISL_691117\|2020-04 | .................... | .................... | ..................... | .................... | .................... | .................... |
| V | 7860 | hCoV-19/Japan/PG-2996/2020\|EPI_ISL_691120\|2020-04 | .................... | .................... | ..................... | .................... | .................... | .................... |
| V | 7861 | hCoV-19/Japan/PG-4090/2020\|EPI_ISL_692331\|2020-07 | .................... | .................... | ..................... | .................... | .................... | .................... |
| V | 7862 | hCoV-19/Japan/PG-4091/2020\|EPI_ISL_692332\|2020-07 | .................... | .................... | ..................... | .................... | .................... | .................... |
| V | 7863 | hCoV-19/Japan/PG-4092/2020\|EPI_ISL_692333\|2020-07 | .................... | .................... | ..................... | .................... | .................... | .................... |
| V | 7864 | hCoV-19/Japan/PG-4093/2020\|EPI_ISL_692334\|2020-07 | .................... | .................... | ..................... | .................... | .................... | .................... |
| V | 7865 | hCoV-19/Japan/PG-4094/2020\|EPI_ISL_692335\|2020-07 | .................... | .................... | ..................... | .................... | .................... | .................... |
| V | 7866 | hCoV-19/Japan/PG-4095/2020\|EPI_ISL_692336\|2020-07 | .................... | .................... | ..................... | .................... | .................... | .................... |
| V | 7867 | hCoV-19/Japan/PG-4096/2020\|EPI_ISL_692337\|2020-07 | .................... | .................... | ..................... | .................... | .................... | .................... |
| V | 7868 | hCoV-19/Japan/PG-4097/2020\|EPI_ISL_692338\|2020-07 | .................... | .................... | ..................... | .................... | .................... | .................... |
| V | 7869 | hCoV-19/Japan/PG-4098/2020\|EPI_ISL_692339\|2020-07 | .................... | .................... | ..................... | .................... | .................... | .................... |
| V | 7870 | hCoV-19/Japan/PG-4099/2020\|EPI_ISL_692340\|2020-07 | .................... | .................... | ..................... | .................... | .................... | .................... |
| V | 7871 | hCoV-19/Japan/PG-4100/2020\|EPI_ISL_692341\|2020-07 | .................... | .................... | ..................... | .................... | .................... | .................... |
| V | 7872 | hCoV-19/Japan/PG-4101/2020\|EPI_ISL_692342\|2020-06 | .................... | .................... | ..................... | .................... | .................... | .................... |
| V | 7873 | hCoV-19/Japan/PG-4102/2020\|EPI_ISL_692343\|2020-06 | .................... | .................... | ..................... | .................... | .................... | .................... |
| V | 7874 | hCoV-19/Japan/PG-4103/2020\|EPI_ISL_692344\|2020-06 | .................... | .................... | ..................... | .................... | .................... | .................... |
| V | 7875 | hCoV-19/Japan/PG-4104/2020\|EPI_ISL_692345\|2020-06 | .................... | .................... | ..................... | .................... | .................... | .................... |
| V | 7876 | hCoV-19/Japan/PG-4105/2020\|EPI_ISL_692346\|2020-06 | .................... | .................... | ..................... | .................... | .................... | .................... |
| V | 7877 | hCoV-19/Japan/PG-4107/2020\|EPI_ISL_692347\|2020-06 | .................... | .................... | ..................... | .................... | .................... | .................... |
| V | 7878 | hCoV-19/Japan/PG-4108/2020\|EPI_ISL_692348\|2020-06 | .................... | .................... | ..................... | .................... | .................... | .................... |
| V | 7879 | hCoV-19/Japan/PG-4109/2020\|EPI_ISL_692349\|2020-06 | .................... | .................... | ..................... | .................... | .................... | .................... |
| V | 7880 | hCoV-19/Japan/PG-4110/2020\|EPI_ISL_692350\|2020-06 | .................... | .................... | ..................... | .................... | .................... | .................... |
| V | 7881 | hCoV-19/Japan/PG-4111/2020\|EPI_ISL_692351\|2020-07 | .................... | .................... | ..................... | .................... | .................... | .................... |
| V | 7883 | hCoV-19/Japan/PG-4113/2020\|EPI_ISL_692353\|2020-07 | .................... | .................... | ..................... | .................... | .................... | .................... |
| V | 7884 | hCoV-19/Japan/PG-4114/2020\|EPI_ISL_692354\|2020-07 | .................... | .................... | ..................... | .................... | .................... | .................... |
| V | 7885 | hCoV-19/Japan/PG-4115/2020\|EPI_ISL_692355\|2020-07 | .................... | .................... | ..................... | .................... | .................... | .................... |
| V | 7886 | hCoV-19/Japan/PG-4116/2020\|EPI_ISL_692356\|2020-07 | .................... | .................... | ..................... | .................... | .................... | .................... |
| V | 7887 | hCoV-19/Japan/PG-4117/2020\|EPI_ISL_692357\|2020-07 | .................... | .................... | ..................... | .................... | .................... | .................... |
| V | 7888 | hCoV-19/Japan/PG-4118/2020\|EPI_ISL_692358\|2020-07 | .................... | .................... | ..................... | .................... | .................... | .................... |
| V | 7889 | hCoV-19/Japan/PG-4119/2020\|EPI_ISL_692359\|2020-07 | .................... | .................... | ..................... | .................... | .................... | .................... |
| V | 7890 | hCoV-19/Japan/PG-4120/2020\|EPI_ISL_692360\|2020-07 | .................... | .................... | ..................... | .................... | .................... | .................... |
| V | 7891 | hCoV-19/Japan/PG-4121/2020\|EPI_ISL_692361\|2020-07 | .................... | .................... | ..................... | .................... | .................... | .................... |
| V | 7892 | hCoV-19/Japan/PG-4122/2020\|EPI_ISL_692362\|2020-07 | .................... | .................... | ..................... | .................... | .................... | .................... |
| V | 7893 | hCoV-19/Japan/PG-4123/2020\|EPI_ISL_692363\|2020-07 | .................... | .................... | ..................... | .................... | .................... | .................... |
| V | 7894 | hCoV-19/Japan/PG-4125/2020\|EPI_ISL_692364\|2020-07 | .................... | .................... | ..................... | .................... | .................... | .................... |
| V | 7895 | hCoV-19/Japan/PG-4126/2020\|EPI_ISL_692365\|2020-07 | .................... | .................... | ..................... | .................... | .................... | .................... |
| V | 7896 | hCoV-19/Japan/PG-4127/2020\|EPI_ISL_692366\|2020-07 | .................... | .................... | ..................... | .................... | .................... | .................... |
| V | 7897 | hCoV-19/Japan/PG-4128/2020\|EPI_ISL_692367\|2020-07 | .................... | .................... | ..................... | .................... | .................... | .................... |
| V | 7898 | hCoV-19/Japan/PG-4129/2020\|EPI_ISL_692368\|2020-07 | .................... | .................... | ..................... | .................... | .................... | .................... |
| V | 7899 | hCoV-19/Japan/PG-4207/2020\|EPI_ISL_692445\|2020-07 | .................... | .................... | ..................... | .................... | .................... | .................... |
| V | 7900 | hCoV-19/Japan/PG-4208/2020\|EPI_ISL_692446\|2020-07 | .................... | .................... | ..................... | .................... | .................... | .................... |

V 7901 hCoV-19/Japan/PG-4209/2020|EPI_ISL_692447|2020-07 .................... .................... .................... .................... .................... ....................
V 7902 hCoV-19/Japan/PG-4210/2020|EPI_ISL_692448|2020-07 .................... .................... .................... .................... .................... ....................
V 7903 hCoV-19/Japan/PG-4212/2020|EPI_ISL_692450|2020-07 .................... .................... .................... .................... .................... ....................
V 7904 hCoV-19/Japan/PG-4213/2020|EPI_ISL_692451|2020-07 .................... .................... .................... .................... .................... ....................
V 7905 hCoV-19/Czech Republic/NRL-10743/2020|EPI_ISL_6936 .................... .................... .................... .................... .................... ....................
V 7906 hCoV-19/Czech Republic/NRL-10473/2020|EPI_ISL_6936 .................... .................... .................... .................... .................... ....................
V 7907 hCoV-19/Czech Republic/NRL-10718/2020|EPI_ISL_6936 .................... .................... .................... .................... .................... ....................
V 7908 hCoV-19/Czech Republic/NRL-10717/2020|EPI_ISL_6936 .................... .................... .................... .................... .................... ....................
V 7909 hCoV-19/Czech Republic/NRL-10560/2020|EPI_ISL_6936 .................... .................... .................... .................... .................... ....................
V 7910 hCoV-19/Czech Republic/NRL-10746/2020|EPI_ISL_6936 .................... .................... .................... .................... .................... ....................
V 7911 hCoV-19/Czech Republic/NRL-10720/2020|EPI_ISL_6936 .................... .................... .................... .................... .................... ....................
V 7912 hCoV-19/Czech Republic/NRL-10552/2020|EPI_ISL_6936 .................... .................... .................... .................... .................... ....................
V 7913 hCoV-19/Czech Republic/NRL-10582/2020|EPI_ISL_6936 .................... .................... .................... .................... .................... ....................
V 7914 hCoV-19/Czech Republic/NRL-10716/2020|EPI_ISL_6936 .................... .................... .................... .................... .................... ....................
V 7915 hCoV-19/Czech Republic/NRL-10503/2020|EPI_ISL_6936 .................... .................... .................... .................... .................... ....................
V 7916 hCoV-19/Czech Republic/NRL-10744/2020|EPI_ISL_6936 .................... .................... .................... .................... .................... ....................
V 7917 hCoV-19/Czech Republic/NRL-10733/2020|EPI_ISL_6936 .................... .................... .................... .................... .................... ....................
V 7918 hCoV-19/Czech Republic/NRL-10565/2020|EPI_ISL_6936 .................... .................... .................... .................... .................... ....................
V 7919 hCoV-19/Czech Republic/NRL-10739/2020|EPI_ISL_6936 .................... .................... .................... .................... .................... ....................
V 7920 hCoV-19/Czech Republic/NRL-10712/2020|EPI_ISL_6936 .................... .................... .................... .................... .................... ....................
V 7921 hCoV-19/Czech Republic/NRL-10645/2020|EPI_ISL_6936 .................... .................... .................... .................... .................... ....................
V 7922 hCoV-19/Czech Republic/NRL-10598/2020|EPI_ISL_6936 .................... .................... .................... .................... .................... ....................
V 7923 hCoV-19/Czech Republic/NRL-10549/2020|EPI_ISL_6936 .................... .................... .................... .................... .................... ....................
V 7924 hCoV-19/Czech Republic/NRL-10710/2020|EPI_ISL_6936 .................... ......C............. .................... .................... .................... ....................
V 7925 hCoV-19/Czech Republic/NRL-10515/2020|EPI_ISL_6936 .................... .................... .................... .................... .................... ....................
V 7926 hCoV-19/Czech Republic/NRL-10715/2020|EPI_ISL_6936 .................... .................... .................... .................... .................... ....................
V 7927 hCoV-19/Czech Republic/NRL-10736/2020|EPI_ISL_6936 .................... .................... .................... .................... .................... ....................
V 7928 hCoV-19/Czech Republic/NRL-10676/2020|EPI_ISL_6936 .................... .................... .................... .................... .................... ....................
V 7929 hCoV-19/Czech Republic/NRL-10487/2020|EPI_ISL_6936 .................... .................... .................... .................... .................... ....................
V 7930 hCoV-19/Switzerland/BL-ETHZ-350001/2020|EPI_ISL_69 .................... .................... .................... .................... .................... ....................
V 7931 hCoV-19/Switzerland/BL-ETHZ-350002/2020|EPI_ISL_69 .................... .................... .................... .................... .................... ....................
V 7932 hCoV-19/Switzerland/JU-ETHZ-350003/2020|EPI_ISL_69 .................... .................... .................... .................... .................... ....................
V 7933 hCoV-19/Switzerland/GE-ETHZ-350004/2020|EPI_ISL_69 .................... .................... .................... .................... .................... ....................
V 7934 hCoV-19/Switzerland/JU-ETHZ-350005/2020|EPI_ISL_69 .................... .................... .................... .................... .................... ....................
V 7935 hCoV-19/Switzerland/VS-ETHZ-350007/2020|EPI_ISL_69 .................... .................... .................... .................... .................... ....................
V 7936 hCoV-19/Switzerland/BL-ETHZ-350008/2020|EPI_ISL_69 .................... .................... .................... .................... .................... ....................
V 7937 hCoV-19/Japan/PG-1554/2020|EPI_ISL_685160|2020-04 .................... .................... .................... .................... .................... ....................
V 7938 hCoV-19/Japan/PG-1555/2020|EPI_ISL_685161|2020-04 .................... .................... .................... .................... .................... ....................
V 7939 hCoV-19/Japan/PG-1556/2020|EPI_ISL_685162|2020-04 .................... .................... .................... .................... .................... ....................
V 7940 hCoV-19/Japan/PG-1557/2020|EPI_ISL_685163|2020-04 .................... .................... .................... .................... .................... ....................
V 7941 hCoV-19/Japan/PG-1558/2020|EPI_ISL_685164|2020-04 .................... .................... .................... .................... .................... ....................
V 7942 hCoV-19/Japan/PG-1559/2020|EPI_ISL_685165|2020-04 .................... .................... .................... .................... .................... ....................
V 7943 hCoV-19/Japan/PG-1560/2020|EPI_ISL_685166|2020-04 .................... .................... .................... .................... .................... ....................
V 7944 hCoV-19/Japan/PG-1561/2020|EPI_ISL_685167|2020-04 .................... .................... .................... .................... .................... ....................
V 7945 hCoV-19/Japan/PG-1562/2020|EPI_ISL_685168|2020-04 .................... .................... .................... .................... .................... ....................
V 7946 hCoV-19/Japan/PG-1563/2020|EPI_ISL_685169|2020-04 .................... .................... .................... .................... .................... ....................
V 7947 hCoV-19/Japan/PG-1564/2020|EPI_ISL_685170|2020-04 .................... .................... .................... .................... .................... ....................
V 7948 hCoV-19/Japan/PG-1565/2020|EPI_ISL_685171|2020-04 .................... .................... .................... .................... .................... ....................
V 7949 hCoV-19/Japan/PG-1566/2020|EPI_ISL_685172|2020-04 .................... .................... .................... .................... .................... ....................
V 7950 hCoV-19/Japan/PG-1567/2020|EPI_ISL_685173|2020-04 .................... .................... .................... .................... .................... ....................
V 7951 hCoV-19/Japan/PG-1568/2020|EPI_ISL_685174|2020-04 .................... .................... .................... .................... .................... ....................
V 7952 hCoV-19/Japan/PG-1569/2020|EPI_ISL_685175|2020-04 .................... .................... .................... .................... .................... ....................
V 7953 hCoV-19/Japan/PG-1570/2020|EPI_ISL_685176|2020-04 .................... .................... .................... .................... .................... ....................
V 7954 hCoV-19/Japan/PG-1571/2020|EPI_ISL_685177|2020-04 .................... .................... .................... .................... .................... ....................
V 7955 hCoV-19/Japan/PG-1572/2020|EPI_ISL_685178|2020-04 .................... .................... .................... .................... .................... ....................
V 7956 hCoV-19/Japan/PG-1573/2020|EPI_ISL_685179|2020-04 .................... .................... .................... .................... .................... ....................
V 7957 hCoV-19/Japan/PG-1574/2020|EPI_ISL_685180|2020-04 .................... .................... .................... .................... .................... ....................
V 7958 hCoV-19/Japan/PG-1575/2020|EPI_ISL_685181|2020-04 .................... .................... .................... .................... .................... ....................
V 7959 hCoV-19/Japan/PG-1576/2020|EPI_ISL_685182|2020-04 .................... .................... .................... .................... .................... ....................
V 7960 hCoV-19/Japan/PG-1577/2020|EPI_ISL_685183|2020-04 .................... .................... .................... .................... .................... ....................
V 7961 hCoV-19/Japan/PG-1578/2020|EPI_ISL_685184|2020-04 .................... .................... .................... .................... .................... ....................
V 7962 hCoV-19/Japan/PG-1579/2020|EPI_ISL_685185|2020-04 .................... .................... .................... .................... .................... ....................
V 7963 hCoV-19/Japan/PG-1580/2020|EPI_ISL_685186|2020-04 .................... .................... .................... .................... .................... ....................
V 7964 hCoV-19/Japan/PG-1581/2020|EPI_ISL_685187|2020-04 .................... .................... .................... .................... .................... ....................
V 7965 hCoV-19/Japan/PG-1582/2020|EPI_ISL_685188|2020-04 .................... .................... .................... .................... .................... ....................
V 7966 hCoV-19/Japan/PG-1583/2020|EPI_ISL_685189|2020-04 .................... .................... .................... .................... .................... ....................
V 7967 hCoV-19/Japan/PG-1584/2020|EPI_ISL_685190|2020-04 .................... .................... .................... .................... .................... ....................
V 7968 hCoV-19/Japan/PG-1585/2020|EPI_ISL_685191|2020-04 .................... .................... .................... .................... .................... ....................
V 7969 hCoV-19/Japan/PG-1586/2020|EPI_ISL_685192|2020-04 .................... .................... .................... .................... .................... ....................
V 7970 hCoV-19/Japan/PG-1587/2020|EPI_ISL_685193|2020-04 .................... .................... .................... .................... .................... ....................
V 7971 hCoV-19/Japan/PG-1588/2020|EPI_ISL_685194|2020-04 .................... .................... .................... .................... .................... ....................
V 7972 hCoV-19/Japan/PG-1589/2020|EPI_ISL_685195|2020-04 .................... .................... .................... .................... .................... ....................
V 7973 hCoV-19/Japan/PG-1590/2020|EPI_ISL_685196|2020-04 .................... .................... .................... .................... .................... ....................
V 7974 hCoV-19/Japan/PG-1591/2020|EPI_ISL_685197|2020-04 .................... .................... .................... .................... .................... ....................
V 7975 hCoV-19/Japan/PG-1592/2020|EPI_ISL_685198|2020-04 .................... .................... .................... .................... .................... ....................
V 7976 hCoV-19/Japan/PG-1593/2020|EPI_ISL_685199|2020-04 .................... .................... .................... .................... .................... ....................
V 7977 hCoV-19/Japan/PG-1594/2020|EPI_ISL_685200|2020-04 .................... .................... .................... .................... .................... ....................
V 7978 hCoV-19/Japan/PG-1595/2020|EPI_ISL_685201|2020-04 .................... .................... .................... .................... .................... ....................
V 7979 hCoV-19/Japan/PG-1596/2020|EPI_ISL_685202|2020-04 .................... .................... .................... .................... .................... ....................
V 7980 hCoV-19/Japan/PG-1597/2020|EPI_ISL_685203|2020-03 .................... .................... .................... .................... .................... ....................

| | | | | | | | | |
|---|---|---|---|---|---|---|---|---|
| V | 7981 | hCoV-19/Japan/PG-1598/2020\|EPI_ISL_685204\|2020-04 | .................... | .................... | .................... | .................... | .................... | .................... |
| V | 7982 | hCoV-19/Japan/PG-1599/2020\|EPI_ISL_685205\|2020-04 | .................... | .................... | .................... | .................... | .................... | .................... |
| V | 7983 | hCoV-19/Japan/PG-1600/2020\|EPI_ISL_685206\|2020-04 | .................... | .................... | .................... | .................... | .................... | .................... |
| V | 7984 | hCoV-19/Japan/PG-1601/2020\|EPI_ISL_685207\|2020-04 | .................... | .................... | .................... | .................... | .................... | .................... |
| V | 7985 | hCoV-19/Japan/PG-1602/2020\|EPI_ISL_685208\|2020-04 | .................... | .................... | .................... | .................... | .................... | .................... |
| V | 7986 | hCoV-19/Japan/PG-1603/2020\|EPI_ISL_685209\|2020-04 | .................... | .................... | .................... | .................... | .................... | .................... |
| V | 7987 | hCoV-19/Japan/PG-8944/2020\|EPI_ISL_686490\|2020-08 | .................... | .................... | .................... | .................... | .................... | .................... |
| V | 7988 | hCoV-19/Japan/PG-8945/2020\|EPI_ISL_686491\|2020-08 | .................... | .................... | .................... | .................... | .................... | .................... |
| V | 7989 | hCoV-19/Japan/PG-8946/2020\|EPI_ISL_686492\|2020-08 | .................... | .................... | .................... | .................... | .................... | .................... |
| V | 7990 | hCoV-19/Japan/PG-8947/2020\|EPI_ISL_686493\|2020-08 | .................... | .................... | .................... | .................... | .................... | .................... |
| V | 7991 | hCoV-19/Japan/PG-8948/2020\|EPI_ISL_686494\|2020-09 | .................... | .................... | .................... | .................... | .................... | .................... |
| V | 7992 | hCoV-19/Japan/PG-8949/2020\|EPI_ISL_686495\|2020-09 | .................... | .................... | .................... | .................... | .................... | .................... |
| V | 7993 | hCoV-19/Japan/PG-8950/2020\|EPI_ISL_686496\|2020-09 | .................... | .................... | .................... | .................... | .................... | .................... |
| V | 7994 | hCoV-19/Japan/PG-8951/2020\|EPI_ISL_686497\|2020-09 | .................... | .................... | .................... | .................... | .................... | .................... |
| V | 7995 | hCoV-19/Japan/PG-8952/2020\|EPI_ISL_686498\|2020-09 | .................... | .................... | .................... | .................... | .................... | .................... |
| V | 7996 | hCoV-19/Japan/PG-8953/2020\|EPI_ISL_686499\|2020-09 | .................... | .................... | .................... | .................... | .................... | .................... |
| V | 7997 | hCoV-19/Japan/PG-8954/2020\|EPI_ISL_686500\|2020-09 | .................... | .................... | .................... | .................... | .................... | .................... |
| V | 7998 | hCoV-19/Japan/PG-8955/2020\|EPI_ISL_686501\|2020-09 | .................... | .................... | .................... | .................... | .................... | .................... |
| V | 7999 | hCoV-19/Japan/PG-8956/2020\|EPI_ISL_686502\|2020-09 | .................... | .................... | .................... | .................... | .................... | .................... |
| V | 8000 | hCoV-19/Japan/PG-8957/2020\|EPI_ISL_686503\|2020-09 | .................... | .................... | .................... | .................... | .................... | .................... |
| V | 8001 | hCoV-19/Japan/PG-8958/2020\|EPI_ISL_686504\|2020-09 | .................... | .................... | .................... | .................... | .................... | .................... |
| V | 8002 | hCoV-19/Japan/PG-8959/2020\|EPI_ISL_686505\|2020-09 | .................... | .................... | .................... | .................... | .................... | .................... |
| V | 8003 | hCoV-19/Japan/PG-8960/2020\|EPI_ISL_686506\|2020-09 | .................... | .................... | .................... | .................... | .................... | .................... |
| V | 8004 | hCoV-19/Japan/PG-8961/2020\|EPI_ISL_686507\|2020-09 | .................... | .................... | .................... | .................... | .................... | .................... |
| V | 8005 | hCoV-19/Japan/PG-8962/2020\|EPI_ISL_686508\|2020-09 | .................... | .................... | .................... | .................... | .................... | .................... |
| V | 8006 | hCoV-19/Japan/PG-8963/2020\|EPI_ISL_686509\|2020-09 | .................... | .................... | .................... | .................... | .................... | .................... |
| V | 8007 | hCoV-19/Japan/PG-8964/2020\|EPI_ISL_686510\|2020-09 | .................... | .................... | .................... | .................... | .................... | .................... |
| V | 8008 | hCoV-19/Japan/PG-8965/2020\|EPI_ISL_686511\|2020-09 | .................... | .................... | .................... | .................... | .................... | .................... |
| V | 8009 | hCoV-19/Japan/PG-8966/2020\|EPI_ISL_686512\|2020-09 | .................... | .................... | .................... | .................... | .................... | .................... |
| V | 8010 | hCoV-19/Japan/PG-8967/2020\|EPI_ISL_686513\|2020-09 | .................... | .................... | .................... | .................... | .................... | .................... |
| V | 8011 | hCoV-19/Japan/PG-8968/2020\|EPI_ISL_686514\|2020-09 | .................... | .................... | .................... | .................... | .................... | .................... |
| V | 8012 | hCoV-19/Japan/PG-8970/2020\|EPI_ISL_686515\|2020-09 | .................... | .................... | .................... | .................... | .................... | .................... |
| V | 8013 | hCoV-19/Japan/PG-8971/2020\|EPI_ISL_686516\|2020-09 | .................... | .................... | .................... | .................... | .................... | .................... |
| V | 8014 | hCoV-19/Japan/PG-8972/2020\|EPI_ISL_686517\|2020-09 | .................... | .................... | .................... | .................... | .................... | .................... |
| V | 8015 | hCoV-19/Japan/PG-8973/2020\|EPI_ISL_686518\|2020-09 | .................... | .................... | .................... | .................... | .................... | .................... |
| V | 8016 | hCoV-19/Japan/PG-8974/2020\|EPI_ISL_686519\|2020-09 | .................... | .................... | .................... | .................... | .................... | .................... |
| V | 8017 | hCoV-19/Japan/PG-8977/2020\|EPI_ISL_686520\|2020-09 | .................... | .................... | .................... | .................... | .................... | .................... |
| V | 8018 | hCoV-19/Japan/PG-8979/2020\|EPI_ISL_686521\|2020-08 | .................... | .................... | .................... | .................... | .................... | .................... |
| V | 8019 | hCoV-19/Japan/PG-8983/2020\|EPI_ISL_686522\|2020-09 | .................... | .................... | .................... | .................... | .................... | .................... |
| V | 8020 | hCoV-19/Japan/PG-8985/2020\|EPI_ISL_686523\|2020-09 | .................... | .................... | .................... | .................... | .................... | .................... |
| V | 8021 | hCoV-19/Japan/PG-8986/2020\|EPI_ISL_686524\|2020-09 | .................... | .................... | .................... | .................... | .................... | .................... |
| V | 8022 | hCoV-19/Japan/PG-8987/2020\|EPI_ISL_686525\|2020-09 | .................... | .................... | .................... | .................... | .................... | .................... |
| V | 8023 | hCoV-19/Japan/PG-8988/2020\|EPI_ISL_686526\|2020-09 | .................... | .................... | .................... | .................... | .................... | .................... |
| V | 8024 | hCoV-19/Japan/PG-8989/2020\|EPI_ISL_686527\|2020-09 | .................... | .................... | .................... | .................... | .................... | .................... |
| V | 8025 | hCoV-19/Japan/PG-8990/2020\|EPI_ISL_686528\|2020-09 | .................... | .................... | .................... | .................... | .................... | .................... |
| V | 8026 | hCoV-19/Japan/PG-8992/2020\|EPI_ISL_686529\|2020-09 | .................... | .................... | .................... | .................... | .................... | .................... |
| V | 8027 | hCoV-19/Japan/PG-8995/2020\|EPI_ISL_686530\|2020-09 | .................... | .................... | .................... | .................... | .................... | .................... |
| V | 8028 | hCoV-19/Japan/PG-8997/2020\|EPI_ISL_686531\|2020-09 | .................... | .................... | .................... | .................... | .................... | .................... |
| V | 8029 | hCoV-19/Japan/PG-8998/2020\|EPI_ISL_686532\|2020-09 | .................... | .................... | .................... | .................... | .................... | .................... |
| V | 8030 | hCoV-19/Japan/PG-8999/2020\|EPI_ISL_686533\|2020-09 | .................... | .................... | .................... | .................... | .................... | .................... |
| V | 8031 | hCoV-19/Japan/PG-9000/2020\|EPI_ISL_686534\|2020-09 | .................... | .................... | .................... | .................... | .................... | .................... |
| V | 8032 | hCoV-19/England/204021372/2020\|EPI_ISL_686538\|2020 | .................... | .................... | .................... | .................... | .................... | .................... |
| V | 8033 | hCoV-19/England/204021577/2020\|EPI_ISL_686539\|2020 | .................... | .................... | .................... | .................... | .................... | .................... |
| V | 8034 | hCoV-19/Japan/PG-6710/2020\|EPI_ISL_687821\|2020-07 | .................... | .................... | .................... | .................... | .................... | .................... |
| V | 8035 | hCoV-19/Japan/PG-6711/2020\|EPI_ISL_687822\|2020-07 | .................... | .................... | .................... | .................... | .................... | .................... |
| V | 8036 | hCoV-19/Japan/PG-6712/2020\|EPI_ISL_687823\|2020-07 | .................... | .................... | .................... | .................... | .................... | .................... |
| V | 8037 | hCoV-19/Japan/PG-6713/2020\|EPI_ISL_687824\|2020-07 | .................... | .................... | .................... | .................... | .................... | .................... |
| V | 8038 | hCoV-19/Japan/PG-6714/2020\|EPI_ISL_687825\|2020-07 | .................... | .................... | .................... | .................... | .................... | .................... |
| V | 8039 | hCoV-19/Japan/PG-6715/2020\|EPI_ISL_687826\|2020-07 | .................... | .................... | .................... | .................... | .................... | .................... |
| V | 8040 | hCoV-19/Japan/PG-6716/2020\|EPI_ISL_687827\|2020-07 | .................... | .................... | .................... | .................... | .................... | .................... |
| V | 8041 | hCoV-19/Japan/PG-6717/2020\|EPI_ISL_687828\|2020-08 | .................... | .................... | .................... | .................... | .................... | .................... |
| V | 8042 | hCoV-19/Japan/PG-6718/2020\|EPI_ISL_687829\|2020-08 | .................... | .................... | .................... | .................... | .................... | .................... |
| V | 8043 | hCoV-19/Japan/PG-6719/2020\|EPI_ISL_687830\|2020-08 | .................... | .................... | .................... | .................... | .................... | .................... |
| V | 8044 | hCoV-19/Japan/PG-6720/2020\|EPI_ISL_687831\|2020-08 | .................... | .................... | .................... | .................... | .................... | .................... |
| V | 8045 | hCoV-19/Japan/PG-6721/2020\|EPI_ISL_687832\|2020-08 | .................... | .................... | .................... | .................... | .................... | .................... |
| V | 8046 | hCoV-19/Japan/PG-6722/2020\|EPI_ISL_687833\|2020-08 | .................... | .................... | .................... | .................... | .................... | .................... |
| V | 8047 | hCoV-19/Japan/PG-6723/2020\|EPI_ISL_687834\|2020-08 | .................... | .................... | .................... | .................... | .................... | .................... |
| V | 8048 | hCoV-19/Japan/PG-6724/2020\|EPI_ISL_687835\|2020-08 | .................... | .................... | .................... | .................... | .................... | .................... |
| V | 8049 | hCoV-19/Japan/PG-6725/2020\|EPI_ISL_687836\|2020-08 | .................... | .................... | .................... | .................... | .................... | .................... |
| V | 8050 | hCoV-19/Japan/PG-6726/2020\|EPI_ISL_687837\|2020-08 | .................... | .................... | .................... | .................... | .................... | .................... |
| V | 8051 | hCoV-19/Japan/PG-6727/2020\|EPI_ISL_687838\|2020-08 | .................... | .................... | .................... | .................... | .................... | .................... |
| V | 8052 | hCoV-19/Japan/PG-6728/2020\|EPI_ISL_687839\|2020-08 | .................... | .................... | .................... | .................... | .................... | .................... |
| V | 8053 | hCoV-19/Japan/PG-6729/2020\|EPI_ISL_687840\|2020-08 | .................... | .................... | .................... | .................... | .................... | .................... |
| V | 8054 | hCoV-19/Japan/PG-6730/2020\|EPI_ISL_687841\|2020-08 | .................... | .................... | .................... | .................... | .................... | .................... |
| V | 8055 | hCoV-19/Japan/PG-6731/2020\|EPI_ISL_687842\|2020-08 | .................... | .................... | .................... | .................... | .................... | .................... |
| V | 8056 | hCoV-19/Japan/PG-6732/2020\|EPI_ISL_687843\|2020-08 | .................... | .................... | .................... | .................... | .................... | .................... |
| V | 8057 | hCoV-19/Japan/PG-6733/2020\|EPI_ISL_687844\|2020-08 | .................... | .................... | .................... | .................... | .................... | .................... |
| V | 8058 | hCoV-19/Japan/PG-6734/2020\|EPI_ISL_687845\|2020-08 | .................... | .................... | .................... | .................... | .................... | .................... |
| V | 8059 | hCoV-19/Japan/PG-6735/2020\|EPI_ISL_687846\|2020-08 | .................... | .................... | .................... | .................... | .................... | .................... |
| V | 8060 | hCoV-19/Japan/PG-6736/2020\|EPI_ISL_687847\|2020-08 | .................... | .................... | .................... | .................... | .................... | .................... |

| | | | | | | | | |
|---|---|---|---|---|---|---|---|---|
| V | 8061 | hCoV-19/Japan/PG-6738/2020\|EPI_ISL_687848\|2020-08 | ···················· | ···················· | ····················· | ···················· | ···················· | ···················· |
| V | 8062 | hCoV-19/Japan/PG-6739/2020\|EPI_ISL_687849\|2020-08 | ···················· | ···················· | ····················· | ···················· | ···················· | ···················· |
| V | 8063 | hCoV-19/Japan/PG-6740/2020\|EPI_ISL_687850\|2020-08 | ···················· | ···················· | ····················· | ···················· | ···················· | ···················· |
| V | 8064 | hCoV-19/Japan/PG-6741/2020\|EPI_ISL_687851\|2020-08 | ···················· | ···················· | ····················· | ···················· | ···················· | ···················· |
| V | 8065 | hCoV-19/Japan/PG-6742/2020\|EPI_ISL_687852\|2020-08 | ···················· | ···················· | ····················· | ···················· | ···················· | ···················· |
| V | 8066 | hCoV-19/Japan/PG-6743/2020\|EPI_ISL_687853\|2020-08 | ···················· | ···················· | ····················· | ···················· | ···················· | ···················· |
| V | 8067 | hCoV-19/Japan/PG-6744/2020\|EPI_ISL_687854\|2020-08 | ···················· | ···················· | ····················· | ···················· | ···················· | ···················· |
| V | 8068 | hCoV-19/Japan/PG-6745/2020\|EPI_ISL_687855\|2020-08 | ···················· | ···················· | ····················· | ···················· | ···················· | ···················· |
| V | 8069 | hCoV-19/Japan/PG-6746/2020\|EPI_ISL_687856\|2020-08 | ···················· | ···················· | ····················· | ···················· | ···················· | ···················· |
| V | 8070 | hCoV-19/Japan/PG-6747/2020\|EPI_ISL_687857\|2020-08 | ···················· | ···················· | ····················· | ···················· | ···················· | ···················· |
| V | 8071 | hCoV-19/Japan/PG-6748/2020\|EPI_ISL_687858\|2020-08 | ···················· | ···················· | ····················· | ···················· | ···················· | ···················· |
| V | 8072 | hCoV-19/Japan/PG-6749/2020\|EPI_ISL_687859\|2020-08 | ···················· | ···················· | ····················· | ···················· | ···················· | ···················· |
| V | 8073 | hCoV-19/Japan/PG-6750/2020\|EPI_ISL_687860\|2020-08 | ···················· | ···················· | ····················· | ···················· | ···················· | ···················· |
| V | 8074 | hCoV-19/Japan/PG-6751/2020\|EPI_ISL_687861\|2020-08 | ···················· | ···················· | ····················· | ···················· | ···················· | ···················· |
| V | 8075 | hCoV-19/Japan/PG-6752/2020\|EPI_ISL_687862\|2020-08 | ···················· | ···················· | ····················· | ···················· | ···················· | ···················· |
| V | 8076 | hCoV-19/Japan/PG-6753/2020\|EPI_ISL_687863\|2020-08 | ···················· | ···················· | ····················· | ···················· | ···················· | ···················· |
| V | 8077 | hCoV-19/Japan/PG-6754/2020\|EPI_ISL_687864\|2020-08 | ···················· | ···················· | ····················· | ···················· | ···················· | ···················· |
| V | 8078 | hCoV-19/Japan/PG-6755/2020\|EPI_ISL_687865\|2020-08 | ···················· | ···················· | ····················· | ···················· | ···················· | ···················· |
| V | 8079 | hCoV-19/Japan/PG-6756/2020\|EPI_ISL_687866\|2020-08 | ···················· | ···················· | ····················· | ···················· | ···················· | ···················· |
| V | 8080 | hCoV-19/Japan/PG-6757/2020\|EPI_ISL_687867\|2020-08 | ···················· | ···················· | ····················· | ···················· | ···················· | ···················· |
| V | 8081 | hCoV-19/Japan/PG-6758/2020\|EPI_ISL_687868\|2020-08 | ···················· | ···················· | ····················· | ···················· | ···················· | ···················· |
| V | 8082 | hCoV-19/Japan/PG-6759/2020\|EPI_ISL_687869\|2020-08 | ···················· | ···················· | ····················· | ···················· | ···················· | ···················· |
| V | 8083 | hCoV-19/Japan/PG-2912/2020\|EPI_ISL_691038\|2020-04 | ···················· | ···················· | ····················· | ···················· | ···················· | ···················· |
| V | 8084 | hCoV-19/Japan/PG-2914/2020\|EPI_ISL_691040\|2020-04 | ···················· | ···················· | ····················· | ···················· | ···················· | ···················· |
| V | 8085 | hCoV-19/Japan/PG-2915/2020\|EPI_ISL_691041\|2020-04 | ···················· | ···················· | ····················· | ···················· | ···················· | ···················· |
| V | 8086 | hCoV-19/Japan/PG-2916/2020\|EPI_ISL_691042\|2020-04 | ···················· | ···················· | ····················· | ···················· | ···················· | ···················· |
| V | 8087 | hCoV-19/Japan/PG-2917/2020\|EPI_ISL_691043\|2020-04 | ···················· | ···················· | ····················· | ···················· | ···················· | ···················· |
| V | 8088 | hCoV-19/Japan/PG-2918/2020\|EPI_ISL_691044\|2020-04 | ···················· | ···················· | ····················· | ···················· | ···················· | ···················· |
| V | 8089 | hCoV-19/Japan/PG-2919/2020\|EPI_ISL_691045\|2020-04 | ···················· | ···················· | ····················· | ···················· | ···················· | ···················· |
| V | 8090 | hCoV-19/Japan/PG-2920/2020\|EPI_ISL_691046\|2020-04 | ···················· | ···················· | ····················· | ···················· | ···················· | ···················· |
| V | 8091 | hCoV-19/Japan/PG-2921/2020\|EPI_ISL_691047\|2020-04 | ···················· | ···················· | ····················· | ···················· | ···················· | ···················· |
| V | 8092 | hCoV-19/Japan/PG-2922/2020\|EPI_ISL_691048\|2020-04 | ···················· | ···················· | ····················· | ···················· | ···················· | ···················· |
| V | 8093 | hCoV-19/Japan/PG-2923/2020\|EPI_ISL_691049\|2020-04 | ···················· | ···················· | ····················· | ···················· | ···················· | ···················· |
| V | 8094 | hCoV-19/Japan/PG-2924/2020\|EPI_ISL_691050\|2020-04 | ···················· | ···················· | ····················· | ···················· | ···················· | ···················· |
| V | 8095 | hCoV-19/Japan/PG-2925/2020\|EPI_ISL_691051\|2020-04 | ···················· | ···················· | ····················· | ···················· | ···················· | ···················· |
| V | 8096 | hCoV-19/Japan/PG-2926/2020\|EPI_ISL_691052\|2020-04 | ···················· | ···················· | ····················· | ···················· | ···················· | ···················· |
| V | 8097 | hCoV-19/Japan/PG-2927/2020\|EPI_ISL_691053\|2020-04 | ···················· | ···················· | ····················· | ···················· | ···················· | ···················· |
| V | 8098 | hCoV-19/Japan/PG-2928/2020\|EPI_ISL_691054\|2020-04 | ···················· | ···················· | ····················· | ···················· | ···················· | ···················· |
| V | 8099 | hCoV-19/Japan/PG-2929/2020\|EPI_ISL_691055\|2020-04 | ···················· | ···················· | ····················· | ···················· | ···················· | ···················· |
| V | 8100 | hCoV-19/Japan/PG-2930/2020\|EPI_ISL_691056\|2020-04 | ···················· | ···················· | ····················· | ···················· | ···················· | ···················· |
| V | 8101 | hCoV-19/Japan/PG-2931/2020\|EPI_ISL_691057\|2020-04 | ···················· | ···················· | ····················· | ···················· | ···················· | ···················· |
| V | 8102 | hCoV-19/Japan/PG-2932/2020\|EPI_ISL_691058\|2020-04 | ···················· | ···················· | ····················· | ···················· | ···················· | ···················· |
| V | 8103 | hCoV-19/Japan/PG-2933/2020\|EPI_ISL_691059\|2020-04 | ···················· | ···················· | ····················· | ···················· | ···················· | ···················· |
| V | 8104 | hCoV-19/Japan/PG-2934/2020\|EPI_ISL_691060\|2020-04 | ···················· | ···················· | ····················· | ···················· | ···················· | ···················· |
| V | 8105 | hCoV-19/Japan/PG-2935/2020\|EPI_ISL_691061\|2020-04 | ···················· | ···················· | ····················· | ···················· | ···················· | ···················· |
| V | 8106 | hCoV-19/Japan/PG-2936/2020\|EPI_ISL_691062\|2020-04 | ···················· | ···················· | ····················· | ···················· | ···················· | ···················· |
| V | 8107 | hCoV-19/Japan/PG-2937/2020\|EPI_ISL_691063\|2020-04 | ···················· | ···················· | ····················· | ···················· | ···················· | ···················· |
| V | 8108 | hCoV-19/Japan/PG-2938/2020\|EPI_ISL_691064\|2020-04 | ···················· | ···················· | ····················· | ···················· | ···················· | ···················· |
| V | 8109 | hCoV-19/Japan/PG-2939/2020\|EPI_ISL_691065\|2020-04 | ···················· | ···················· | ····················· | ···················· | ···················· | ···················· |
| V | 8110 | hCoV-19/Japan/PG-2940/2020\|EPI_ISL_691066\|2020-04 | ···················· | ···················· | ····················· | ···················· | ···················· | ···················· |
| V | 8111 | hCoV-19/Japan/PG-2941/2020\|EPI_ISL_691067\|2020-04 | ···················· | ···················· | ····················· | ···················· | ···················· | ···················· |
| V | 8112 | hCoV-19/Japan/PG-2942/2020\|EPI_ISL_691068\|2020-04 | ···················· | ···················· | ····················· | ···················· | ···················· | ···················· |
| V | 8113 | hCoV-19/Japan/PG-2943/2020\|EPI_ISL_691069\|2020-04 | ···················· | ···················· | ····················· | ···················· | ···················· | ···················· |
| V | 8114 | hCoV-19/Japan/PG-2944/2020\|EPI_ISL_691070\|2020-04 | ···················· | ···················· | ····················· | ···················· | ···················· | ···················· |
| V | 8115 | hCoV-19/Japan/PG-2945/2020\|EPI_ISL_691071\|2020-04 | ···················· | ···················· | ····················· | ···················· | ···················· | ···················· |
| V | 8116 | hCoV-19/Japan/PG-2946/2020\|EPI_ISL_691072\|2020-04 | ···················· | ···················· | ····················· | ···················· | ···················· | ···················· |
| V | 8117 | hCoV-19/Japan/PG-2947/2020\|EPI_ISL_691073\|2020-04 | ···················· | ···················· | ····················· | ···················· | ···················· | ···················· |
| V | 8118 | hCoV-19/Japan/PG-2948/2020\|EPI_ISL_691074\|2020-04 | ···················· | ···················· | ····················· | ···················· | ···················· | ···················· |
| V | 8119 | hCoV-19/Japan/PG-2949/2020\|EPI_ISL_691075\|2020-04 | ···················· | ···················· | ····················· | ···················· | ···················· | ···················· |
| V | 8120 | hCoV-19/Japan/PG-2950/2020\|EPI_ISL_691076\|2020-04 | ···················· | ···················· | ····················· | ···················· | ···················· | ···················· |
| V | 8121 | hCoV-19/Japan/PG-2951/2020\|EPI_ISL_691077\|2020-04 | ···················· | ···················· | ····················· | ···················· | ···················· | ···················· |
| V | 8122 | hCoV-19/Japan/PG-2952/2020\|EPI_ISL_691078\|2020-04 | ···················· | ···················· | ····················· | ···················· | ···················· | ···················· |
| V | 8123 | hCoV-19/Japan/PG-2953/2020\|EPI_ISL_691079\|2020-04 | ···················· | ···················· | ····················· | ···················· | ···················· | ···················· |
| V | 8124 | hCoV-19/Japan/PG-2954/2020\|EPI_ISL_691080\|2020-04 | ···················· | ···················· | ····················· | ···················· | ···················· | ···················· |
| V | 8125 | hCoV-19/Japan/PG-2955/2020\|EPI_ISL_691081\|2020-04 | ···················· | ···················· | ····················· | ···················· | ···················· | ···················· |
| V | 8126 | hCoV-19/Japan/PG-2956/2020\|EPI_ISL_691082\|2020-04 | ···················· | ···················· | ····················· | ···················· | ···················· | ···················· |
| V | 8127 | hCoV-19/Japan/PG-2957/2020\|EPI_ISL_691083\|2020-04 | ···················· | ···················· | ····················· | ···················· | ···················· | ···················· |
| V | 8128 | hCoV-19/Japan/PG-2958/2020\|EPI_ISL_691084\|2020-04 | ···················· | ···················· | ····················· | ···················· | ···················· | ···················· |
| V | 8129 | hCoV-19/Japan/PG-2959/2020\|EPI_ISL_691085\|2020-04 | ···················· | ···················· | ····················· | ···················· | ···················· | ···················· |
| V | 8130 | hCoV-19/Japan/PG-2960/2020\|EPI_ISL_691086\|2020-04 | ···················· | ···················· | ····················· | ···················· | ···················· | ···················· |
| V | 8131 | hCoV-19/Japan/PG-2961/2020\|EPI_ISL_691087\|2020-04 | ···················· | ···················· | ····················· | ···················· | ···················· | ···················· |
| V | 8132 | hCoV-19/Japan/PG-4130/2020\|EPI_ISL_692369\|2020-07 | ···················· | ···················· | ····················· | ···················· | ···················· | ···················· |
| V | 8133 | hCoV-19/Japan/PG-4131/2020\|EPI_ISL_692370\|2020-07 | ···················· | ···················· | ····················· | ···················· | ···················· | ···················· |
| V | 8134 | hCoV-19/Japan/PG-4132/2020\|EPI_ISL_692371\|2020-07 | ···················· | ···················· | ····················· | ···················· | ···················· | ···················· |
| V | 8135 | hCoV-19/Japan/PG-4133/2020\|EPI_ISL_692372\|2020-07 | ···················· | ···················· | ····················· | ···················· | ···················· | ···················· |
| V | 8136 | hCoV-19/Japan/PG-4134/2020\|EPI_ISL_692373\|2020-07 | ···················· | ···················· | ····················· | ···················· | ···················· | ···················· |
| V | 8137 | hCoV-19/Japan/PG-4135/2020\|EPI_ISL_692374\|2020-07 | ···················· | ···················· | ····················· | ···················· | ···················· | ···················· |
| V | 8138 | hCoV-19/Japan/PG-4136/2020\|EPI_ISL_692375\|2020-07 | ···················· | ···················· | ····················· | ···················· | ···················· | ···················· |
| V | 8139 | hCoV-19/Japan/PG-4137/2020\|EPI_ISL_692376\|2020-07 | ···················· | ···················· | ····················· | ···················· | ···················· | ···················· |
| V | 8140 | hCoV-19/Japan/PG-4138/2020\|EPI_ISL_692377\|2020-07 | ···················· | ·A·················· | ····················· | ···················· | ···················· | ···················· |

| | | | | | | | | |
|---|---|---|---|---|---|---|---|---|
| V | 8142 | hCoV-19/Japan/PG-4140/2020\|EPI_ISL_692379\|2020-07 | .................... | .................... | ..................... | .................... | .................... | .................... |
| V | 8143 | hCoV-19/Japan/PG-4141/2020\|EPI_ISL_692380\|2020-07 | .................... | .................... | ..................... | .................... | .................... | .................... |
| V | 8144 | hCoV-19/Japan/PG-4142/2020\|EPI_ISL_692381\|2020-07 | .................... | .................... | ..................... | .................... | .................... | .................... |
| V | 8145 | hCoV-19/Japan/PG-4143/2020\|EPI_ISL_692382\|2020-07 | .................... | .................... | ..................... | .................... | .................... | .................... |
| V | 8146 | hCoV-19/Japan/PG-4144/2020\|EPI_ISL_692383\|2020-07 | .................... | .................... | ..................... | .................... | .................... | .................... |
| V | 8147 | hCoV-19/Japan/PG-4145/2020\|EPI_ISL_692384\|2020-07 | .................... | .................... | ..................... | .................... | .................... | .................... |
| V | 8148 | hCoV-19/Japan/PG-4146/2020\|EPI_ISL_692385\|2020-07 | .................... | .................... | ..................... | .................... | .................... | .................... |
| V | 8149 | hCoV-19/Japan/PG-4147/2020\|EPI_ISL_692386\|2020-07 | .................... | .................... | ..................... | .................... | .................... | .................... |
| V | 8150 | hCoV-19/Japan/PG-4148/2020\|EPI_ISL_692387\|2020-07 | .................... | .................... | ..................... | .................... | .................... | .................... |
| V | 8151 | hCoV-19/Japan/PG-4149/2020\|EPI_ISL_692388\|2020-07 | .................... | .................... | ..................... | .................... | .................... | .................... |
| V | 8152 | hCoV-19/Japan/PG-4150/2020\|EPI_ISL_692389\|2020-07 | .................... | .................... | ..................... | .................... | .................... | .................... |
| V | 8153 | hCoV-19/Japan/PG-4151/2020\|EPI_ISL_692390\|2020-07 | .................... | .................... | ..................... | .................... | .................... | .................... |
| V | 8154 | hCoV-19/Japan/PG-4152/2020\|EPI_ISL_692391\|2020-07 | .................... | .................... | ..................... | .................... | .................... | .................... |
| V | 8155 | hCoV-19/Japan/PG-4153/2020\|EPI_ISL_692392\|2020-07 | .................... | .................... | ..................... | .................... | .................... | .................... |
| V | 8156 | hCoV-19/Japan/PG-4154/2020\|EPI_ISL_692393\|2020-07 | .................... | .................... | ..................... | .................... | .................... | .................... |
| V | 8157 | hCoV-19/Japan/PG-4155/2020\|EPI_ISL_692394\|2020-07 | .................... | .................... | ..................... | .................... | .................... | .................... |
| V | 8158 | hCoV-19/Japan/PG-4156/2020\|EPI_ISL_692395\|2020-07 | .................... | .................... | ..................... | .................... | .................... | .................... |
| V | 8159 | hCoV-19/Japan/PG-4157/2020\|EPI_ISL_692396\|2020-07 | .................... | .................... | ..................... | .................... | .................... | .................... |
| V | 8160 | hCoV-19/Japan/PG-4158/2020\|EPI_ISL_692397\|2020-07 | .................... | .................... | ..................... | .................... | .................... | .................... |
| V | 8161 | hCoV-19/Japan/PG-4159/2020\|EPI_ISL_692398\|2020-07 | .................... | .................... | ..................... | .................... | .................... | .................... |
| V | 8162 | hCoV-19/Japan/PG-4160/2020\|EPI_ISL_692399\|2020-07 | .................... | .................... | ..................... | .................... | .................... | .................... |
| V | 8166 | hCoV-19/Portugal/PT1599/2020\|EPI_ISL_693603\|2020-0 | .................... | .................... | ..................... | .................... | .................... | .................... |
| V | 8167 | hCoV-19/Portugal/PT1601/2020\|EPI_ISL_693604\|2020-0 | .................... | .................... | ..................... | .................... | .................... | .................... |
| V | 8168 | hCoV-19/Portugal/PT1602/2020\|EPI_ISL_693605\|2020-0 | .................... | .................... | ..................... | .................... | .................... | .................... |
| V | 8172 | hCoV-19/Portugal/PT1606/2020\|EPI_ISL_693609\|2020-0 | .................... | .................... | ..................... | .................... | .................... | .................... |
| V | 8174 | hCoV-19/Portugal/PT1608/2020\|EPI_ISL_693611\|2020-0 | .................... | .................... | ..................... | .................... | .................... | .................... |
| V | 8175 | hCoV-19/Portugal/PT1610/2020\|EPI_ISL_693612\|2020-0 | .................... | .................... | ..................... | .................... | .................... | .................... |
| V | 8177 | hCoV-19/Portugal/PT1612/2020\|EPI_ISL_693614\|2020-0 | .................... | .................... | ..................... | .................... | .................... | .................... |
| V | 8178 | hCoV-19/Portugal/PT1613/2020\|EPI_ISL_693615\|2020-0 | .................... | .................... | ..................... | .................... | .................... | .................... |
| V | 8179 | hCoV-19/Portugal/PT1614/2020\|EPI_ISL_693616\|2020-0 | .................... | .........A.......... | ..................... | .................... | .................... | ....N............... |
| V | 8180 | hCoV-19/Portugal/PT1615/2020\|EPI_ISL_693617\|2020-0 | .................... | .................... | ..................... | .................... | .................... | .................... |
| V | 8181 | hCoV-19/Portugal/PT1616/2020\|EPI_ISL_693618\|2020-0 | .................... | .................... | ..................... | .................... | ..........N......... | .................... |
| V | 8183 | hCoV-19/Portugal/PT1619/2020\|EPI_ISL_693620\|2020-0 | .................... | .................... | ..................... | .................... | .................... | .................... |
| V | 8184 | hCoV-19/Portugal/PT1620/2020\|EPI_ISL_693621\|2020-0 | .................... | .................... | ..................... | .................... | .................... | .................... |
| V | 8185 | hCoV-19/Portugal/PT1621/2020\|EPI_ISL_693622\|2020-0 | .................... | .................... | ..................... | .................... | .................... | .................... |
| V | 8186 | hCoV-19/Portugal/PT1622/2020\|EPI_ISL_693623\|2020-0 | .................... | .................... | ..................... | .................... | .................... | .................... |
| V | 8187 | hCoV-19/Portugal/PT1623/2020\|EPI_ISL_693624\|2020-0 | .................... | .................... | ..................... | .................... | .................... | .................... |
| V | 8188 | hCoV-19/Portugal/PT1624/2020\|EPI_ISL_693625\|2020-0 | .................... | .................... | ..................... | .................... | .................... | .................... |
| V | 8190 | hCoV-19/Portugal/PT1627/2020\|EPI_ISL_693627\|2020-0 | .................... | .................... | ..................... | .................... | .................... | .................... |
| V | 8191 | hCoV-19/Portugal/PT1628/2020\|EPI_ISL_693628\|2020-0 | .................... | .................... | ..................... | .................... | .................... | .................... |
| V | 8192 | hCoV-19/Portugal/PT1629/2020\|EPI_ISL_693629\|2020-0 | .................... | .................... | ..................... | .................... | .................... | .................... |
| V | 8193 | hCoV-19/Japan/PG-1495/2020\|EPI_ISL_685102\|2020-04 | .................... | .................... | ..................... | .................... | .................... | .................... |
| V | 8194 | hCoV-19/Japan/PG-1497/2020\|EPI_ISL_685104\|2020-04 | .................... | .................... | ..................... | .................... | .................... | .................... |
| V | 8195 | hCoV-19/Japan/PG-1498/2020\|EPI_ISL_685105\|2020-04 | .................... | .................... | ..................... | .................... | .................... | .................... |
| V | 8196 | hCoV-19/Japan/PG-1499/2020\|EPI_ISL_685106\|2020-04 | .................... | .................... | ..................... | .................... | .................... | .................... |
| V | 8197 | hCoV-19/Japan/PG-1500/2020\|EPI_ISL_685107\|2020-04 | .................... | .................... | ..................... | .................... | .................... | .................... |
| V | 8198 | hCoV-19/Japan/PG-1501/2020\|EPI_ISL_685108\|2020-04 | .................... | .................... | ..................... | .................... | .................... | .................... |
| V | 8199 | hCoV-19/Japan/PG-1502/2020\|EPI_ISL_685109\|2020-04 | .................... | .................... | ..................... | .................... | .................... | .................... |
| V | 8200 | hCoV-19/Japan/PG-1503/2020\|EPI_ISL_685110\|2020-04 | .................... | .................... | ..................... | .................... | .................... | .................... |
| V | 8201 | hCoV-19/Japan/PG-1504/2020\|EPI_ISL_685111\|2020-04 | .................... | .................... | ..................... | .................... | .................... | .................... |
| V | 8202 | hCoV-19/Japan/PG-1505/2020\|EPI_ISL_685112\|2020-04 | .................... | .................... | ..................... | .................... | .................... | .................... |
| V | 8203 | hCoV-19/Japan/PG-1506/2020\|EPI_ISL_685113\|2020-04 | .................... | .................... | ..................... | .................... | .................... | .................... |
| V | 8204 | hCoV-19/Japan/PG-1507/2020\|EPI_ISL_685114\|2020-04 | .................... | .................... | ..................... | .................... | .................... | .................... |
| V | 8205 | hCoV-19/Japan/PG-1508/2020\|EPI_ISL_685115\|2020-04 | .................... | .................... | ..................... | .................... | .................... | .................... |
| V | 8206 | hCoV-19/Japan/PG-1509/2020\|EPI_ISL_685116\|2020-04 | .................... | .................... | ..................... | .................... | .................... | .................... |
| V | 8207 | hCoV-19/Japan/PG-1510/2020\|EPI_ISL_685117\|2020-04 | .................... | .................... | ..................... | .................... | .................... | .................... |
| V | 8208 | hCoV-19/Japan/PG-1511/2020\|EPI_ISL_685118\|2020-04 | .................... | .................... | ..................... | .................... | .................... | .................... |
| V | 8209 | hCoV-19/Japan/PG-1512/2020\|EPI_ISL_685119\|2020-04 | .................... | .................... | ..................... | .................... | .................... | .................... |
| V | 8210 | hCoV-19/Japan/PG-1513/2020\|EPI_ISL_685120\|2020-04 | .................... | .................... | ..................... | .................... | .................... | .................... |
| V | 8211 | hCoV-19/Japan/PG-1514/2020\|EPI_ISL_685121\|2020-04 | .................... | .................... | ..................... | .................... | .................... | .................... |
| V | 8212 | hCoV-19/Japan/PG-1515/2020\|EPI_ISL_685122\|2020-04 | .................... | .................... | ..................... | .................... | .................... | .................... |
| V | 8213 | hCoV-19/Japan/PG-1516/2020\|EPI_ISL_685123\|2020-04 | .................... | .................... | ..................... | .................... | .................... | .................... |
| V | 8214 | hCoV-19/Japan/PG-1517/2020\|EPI_ISL_685124\|2020-04 | .................... | .................... | ..................... | .................... | .................... | .................... |
| V | 8215 | hCoV-19/Japan/PG-1519/2020\|EPI_ISL_685126\|2020-04 | .................... | .................... | ..................... | .................... | .................... | .................... |
| V | 8216 | hCoV-19/Japan/PG-1520/2020\|EPI_ISL_685127\|2020-04 | .................... | .................... | ..................... | .................... | .................... | .................... |
| V | 8217 | hCoV-19/Japan/PG-1521/2020\|EPI_ISL_685128\|2020-04 | .................... | .................... | ..................... | .................... | .................... | .................... |
| V | 8218 | hCoV-19/Japan/PG-1522/2020\|EPI_ISL_685129\|2020-04 | .................... | .................... | ..................... | .................... | .................... | .................... |
| V | 8219 | hCoV-19/Japan/PG-1523/2020\|EPI_ISL_685130\|2020-04 | .................... | .................... | ..................... | .................... | .................... | .................... |
| V | 8220 | hCoV-19/Japan/PG-1524/2020\|EPI_ISL_685131\|2020-04 | .................... | .................... | ..................... | .................... | .................... | .................... |
| V | 8221 | hCoV-19/Japan/PG-1526/2020\|EPI_ISL_685132\|2020-03 | .................... | .................... | ..................... | .................... | .................... | .................... |
| V | 8222 | hCoV-19/Japan/PG-1527/2020\|EPI_ISL_685133\|2020-03 | .................... | .................... | ..................... | .................... | .................... | .................... |
| V | 8223 | hCoV-19/Japan/PG-1528/2020\|EPI_ISL_685134\|2020-03 | .................... | .................... | ..................... | .................... | .................... | .................... |
| V | 8224 | hCoV-19/Japan/PG-1529/2020\|EPI_ISL_685135\|2020-03 | .................... | .................... | ..................... | .................... | .................... | .................... |
| V | 8225 | hCoV-19/Japan/PG-1530/2020\|EPI_ISL_685136\|2020-04 | .................... | .................... | ..................... | .................... | .................... | .................... |
| V | 8226 | hCoV-19/Japan/PG-1531/2020\|EPI_ISL_685137\|2020-04 | .................... | .................... | ..................... | .................... | .................... | .................... |
| V | 8227 | hCoV-19/Japan/PG-1532/2020\|EPI_ISL_685138\|2020-04 | .................... | .................... | ..................... | .................... | .................... | .................... |
| V | 8228 | hCoV-19/Japan/PG-1533/2020\|EPI_ISL_685139\|2020-04 | .................... | .................... | ..................... | .................... | .................... | .................... |
| V | 8229 | hCoV-19/Japan/PG-1534/2020\|EPI_ISL_685140\|2020-04 | .................... | .................... | ..................... | .................... | .................... | .................... |
| V | 8230 | hCoV-19/Japan/PG-1535/2020\|EPI_ISL_685141\|2020-04 | .................... | .................... | ..................... | .................... | .................... | .................... |
| V | 8231 | hCoV-19/Japan/PG-1536/2020\|EPI_ISL_685142\|2020-04 | .................... | .................... | ..................... | .................... | .................... | .................... |

V 8232 hCoV-19/Japan/PG-1537/2020|EPI_ISL_685143|2020-04 .................... .................... ..................... .................... .................... ....................
V 8234 hCoV-19/Japan/PG-1539/2020|EPI_ISL_685145|2020-04 .................... .................... ..................... .................... .................... ....................
V 8235 hCoV-19/Japan/PG-1540/2020|EPI_ISL_685146|2020-04 .................... .................... ..................... .................... .................... ....................
V 8236 hCoV-19/Japan/PG-1541/2020|EPI_ISL_685147|2020-04 .................... .................... ..................... .................... .................... ....................
V 8237 hCoV-19/Japan/PG-1542/2020|EPI_ISL_685148|2020-04 .................... .................... ..................... .................... .................... ....................
V 8238 hCoV-19/Japan/PG-1543/2020|EPI_ISL_685149|2020-04 .................... .................... ..................... .................... .................... ....................
V 8240 hCoV-19/Japan/PG-1545/2020|EPI_ISL_685151|2020-04 .................... .................... ..................... .................... .................... ....................
V 8241 hCoV-19/Japan/PG-1546/2020|EPI_ISL_685152|2020-04 .................... .................... ..................... .................... .................... ....................
V 8242 hCoV-19/Japan/PG-1547/2020|EPI_ISL_685153|2020-04 .................... .................... ..................... .................... .................... ....................
V 8243 hCoV-19/Japan/PG-1548/2020|EPI_ISL_685154|2020-04 .................... .................... ..................... .................... .................... ....................
V 8244 hCoV-19/Japan/PG-1549/2020|EPI_ISL_685155|2020-04 .................... .................... ..................... .................... .................... ....................
V 8246 hCoV-19/Japan/PG-1551/2020|EPI_ISL_685157|2020-04 .................... .................... ..................... .................... .................... ....................
V 8247 hCoV-19/Japan/PG-1552/2020|EPI_ISL_685158|2020-04 .................... .................... ..................... .................... .................... ....................
V 8248 hCoV-19/Japan/PG-1553/2020|EPI_ISL_685159|2020-04 .................... .................... ..................... .................... .................... ....................
V 8249 hCoV-19/Japan/PG-8887/2020|EPI_ISL_686433|2020-08 .................... .................... ..................... .................... .................... ....................
V 8250 hCoV-19/Japan/PG-8889/2020|EPI_ISL_686435|2020-08 .................... .................... ..................... .................... .................... ....................
V 8251 hCoV-19/Japan/PG-8890/2020|EPI_ISL_686436|2020-08 .................... .................... ..................... .................... .................... ....................
V 8252 hCoV-19/Japan/PG-8891/2020|EPI_ISL_686437|2020-08 .................... .................... ..................... .................... .................... ....................
V 8253 hCoV-19/Japan/PG-8892/2020|EPI_ISL_686438|2020-08 .................... .................... ..................... .................... .................... ....................
V 8254 hCoV-19/Japan/PG-8893/2020|EPI_ISL_686439|2020-08 .................... .................... ..................... .................... .................... ....................
V 8255 hCoV-19/Japan/PG-8894/2020|EPI_ISL_686440|2020-08 .................... .................... ..................... .................... .................... ....................
V 8256 hCoV-19/Japan/PG-8895/2020|EPI_ISL_686441|2020-08 .................... .................... ..................... .................... .................... ....................
V 8257 hCoV-19/Japan/PG-8896/2020|EPI_ISL_686442|2020-08 .................... .................... ..................... .................... .................... ....................
V 8258 hCoV-19/Japan/PG-8897/2020|EPI_ISL_686443|2020-08 .................... .................... ..................... .................... .................... ....................
V 8259 hCoV-19/Japan/PG-8898/2020|EPI_ISL_686444|2020-08 .................... .................... ..................... .................... .................... ....................
V 8260 hCoV-19/Japan/PG-8899/2020|EPI_ISL_686445|2020-08 .................... .................... ..................... .................... .................... ....................
V 8261 hCoV-19/Japan/PG-8900/2020|EPI_ISL_686446|2020-08 .................... .................... ..................... .................... .................... ....................
V 8262 hCoV-19/Japan/PG-8901/2020|EPI_ISL_686447|2020-08 .................... .................... ..................... .................... .................... ....................
V 8263 hCoV-19/Japan/PG-8902/2020|EPI_ISL_686448|2020-09 .................... .................... ..................... .................... .................... ....................
V 8264 hCoV-19/Japan/PG-8903/2020|EPI_ISL_686449|2020-09 .................... .................... ..................... .................... .................... ....................
V 8265 hCoV-19/Japan/PG-8904/2020|EPI_ISL_686450|2020-09 .................... .................... ..................... .................... .................... ....................
V 8266 hCoV-19/Japan/PG-8905/2020|EPI_ISL_686451|2020-09 .................... .................... ..................... .................... .................... ....................
V 8267 hCoV-19/Japan/PG-8906/2020|EPI_ISL_686452|2020-09 .................... .................... ..................... .................... .................... ....................
V 8268 hCoV-19/Japan/PG-8907/2020|EPI_ISL_686453|2020-09 .................... .................... ..................... .................... .................... ....................
V 8269 hCoV-19/Japan/PG-8908/2020|EPI_ISL_686454|2020-09 .................... .................... ..................... .................... .................... ....................
V 8270 hCoV-19/Japan/PG-8909/2020|EPI_ISL_686455|2020-09 .................... .................... ..................... .................... .................... ....................
V 8271 hCoV-19/Japan/PG-8910/2020|EPI_ISL_686456|2020-09 .................... .................... ..................... .................... .................... ....................
V 8272 hCoV-19/Japan/PG-8911/2020|EPI_ISL_686457|2020-09 .................... .................... ..................... .................... .................... ....................
V 8273 hCoV-19/Japan/PG-8912/2020|EPI_ISL_686458|2020-09 .................... .................... ..................... .................... .................... ....................
V 8274 hCoV-19/Japan/PG-8913/2020|EPI_ISL_686459|2020-09 .................... .................... ..................... .................... .................... ....................
V 8275 hCoV-19/Japan/PG-8914/2020|EPI_ISL_686460|2020-09 .................... .................... ..................... .................... .................... ....................
V 8276 hCoV-19/Japan/PG-8915/2020|EPI_ISL_686461|2020-09 .................... .A.................. ..................... .................... .................... ....................
V 8277 hCoV-19/Japan/PG-8916/2020|EPI_ISL_686462|2020-09 .................... .................... ..................... .................... .................... ....................
V 8278 hCoV-19/Japan/PG-8917/2020|EPI_ISL_686463|2020-09 .................... .................... ..................... .................... .................... ....................
V 8279 hCoV-19/Japan/PG-8918/2020|EPI_ISL_686464|2020-09 .................... .................... ..................... .................... .................... ....................
V 8280 hCoV-19/Japan/PG-8919/2020|EPI_ISL_686465|2020-09 .................... .................... ..................... .................... .................... ....................
V 8282 hCoV-19/Japan/PG-8921/2020|EPI_ISL_686467|2020-09 .................... .................... ..................... .................... .................... ....................
V 8283 hCoV-19/Japan/PG-8922/2020|EPI_ISL_686468|2020-09 .................... .................... ..................... .................... .................... ....................
V 8284 hCoV-19/Japan/PG-8923/2020|EPI_ISL_686469|2020-08 .................... .................... ..................... .................... .................... ....................
V 8285 hCoV-19/Japan/PG-8924/2020|EPI_ISL_686470|2020-08 .................... .................... ..................... .................... .................... ....................
V 8286 hCoV-19/Japan/PG-8925/2020|EPI_ISL_686471|2020-08 .................... .................... ..................... .................... .................... ....................
V 8287 hCoV-19/Japan/PG-8926/2020|EPI_ISL_686472|2020-08 .................... .................... ..................... .................... .................... ....................
V 8288 hCoV-19/Japan/PG-8927/2020|EPI_ISL_686473|2020-08 .................... .................... ..................... .................... .................... ....................
V 8289 hCoV-19/Japan/PG-8928/2020|EPI_ISL_686474|2020-08 .................... .................... ..................... .................... .................... ....................
V 8290 hCoV-19/Japan/PG-8929/2020|EPI_ISL_686475|2020-08 .................... .................... ..................... .................... .................... ....................
V 8291 hCoV-19/Japan/PG-8930/2020|EPI_ISL_686476|2020-08 .................... .................... ..................... .................... .................... ....................
V 8292 hCoV-19/Japan/PG-8931/2020|EPI_ISL_686477|2020-08 .................... .................... ..................... .................... .................... ....................
V 8293 hCoV-19/Japan/PG-8932/2020|EPI_ISL_686478|2020-08 .................... .................... ..................... .................... .................... ....................
V 8294 hCoV-19/Japan/PG-8933/2020|EPI_ISL_686479|2020-08 .................... .................... ..................... .................... .................... ....................
V 8295 hCoV-19/Japan/PG-8934/2020|EPI_ISL_686480|2020-08 .................... .................... ..................... .................... .................... ....................
V 8296 hCoV-19/Japan/PG-8935/2020|EPI_ISL_686481|2020-08 .................... .................... ..................... .................... .................... ....................
V 8297 hCoV-19/Japan/PG-8936/2020|EPI_ISL_686482|2020-08 .................... .................... ..................... .................... .................... ....................
V 8298 hCoV-19/Japan/PG-8937/2020|EPI_ISL_686483|2020-08 .................... .................... ..................... .................... .................... ....................
V 8299 hCoV-19/Japan/PG-8938/2020|EPI_ISL_686484|2020-08 .................... .................... ..................... .................... .................... ....................
V 8300 hCoV-19/Japan/PG-8939/2020|EPI_ISL_686485|2020-08 .................... .................... ..................... .................... .................... ....................
V 8301 hCoV-19/Japan/PG-8940/2020|EPI_ISL_686486|2020-08 .................... .................... ..................... .................... .................... ....................
V 8302 hCoV-19/Japan/PG-8941/2020|EPI_ISL_686487|2020-08 .................... .................... ..................... .................... .................... ....................
V 8303 hCoV-19/Japan/PG-8942/2020|EPI_ISL_686488|2020-08 .................... .................... ..................... .................... .................... ....................
V 8304 hCoV-19/Japan/PG-8943/2020|EPI_ISL_686489|2020-08 .................... .................... ..................... .................... .................... ....................
V 8305 hCoV-19/Japan/PG-6652/2020|EPI_ISL_687764|2020-07 .................... .................... ..................... .................... .................... ....................
V 8306 hCoV-19/Japan/PG-6653/2020|EPI_ISL_687765|2020-07 .................... .................... ..................... .................... .................... ....................
V 8307 hCoV-19/Japan/PG-6654/2020|EPI_ISL_687766|2020-07 .................... .................... ..................... .................... .................... ....................
V 8308 hCoV-19/Japan/PG-6655/2020|EPI_ISL_687767|2020-07 .................... .................... ..................... .................... .................... ....................
V 8309 hCoV-19/Japan/PG-6656/2020|EPI_ISL_687768|2020-07 .................... .................... ..................... .................... .................... ....................
V 8310 hCoV-19/Japan/PG-6658/2020|EPI_ISL_687769|2020-07 .................... .................... ..................... .................... .................... ....................
V 8311 hCoV-19/Japan/PG-6659/2020|EPI_ISL_687770|2020-07 .................... .................... ..................... .................... .................... ....................
V 8312 hCoV-19/Japan/PG-6660/2020|EPI_ISL_687771|2020-07 .................... .................... ..................... .................... .................... ....................
V 8313 hCoV-19/Japan/PG-6661/2020|EPI_ISL_687772|2020-07 .................... .................... ..................... .................... .................... ....................
V 8314 hCoV-19/Japan/PG-6662/2020|EPI_ISL_687773|2020-07 .................... .................... ..................... .................... .................... ....................
V 8315 hCoV-19/Japan/PG-6663/2020|EPI_ISL_687774|2020-07 .................... .................... ..................... .................... .................... ....................

| | | | | | | | |
|---|---|---|---|---|---|---|---|
| V | 8316 hCoV-19/Japan/PG-6664/2020\|EPI_ISL_687775\|2020-07 | ···················· | ···················· | ····················· | ···················· | ···················· | ···················· |
| V | 8317 hCoV-19/Japan/PG-6665/2020\|EPI_ISL_687776\|2020-07 | ···················· | ···················· | ····················· | ···················· | ···················· | ···················· |
| V | 8318 hCoV-19/Japan/PG-6666/2020\|EPI_ISL_687777\|2020-07 | ···················· | ···················· | ····················· | ···················· | ···················· | ···················· |
| V | 8319 hCoV-19/Japan/PG-6667/2020\|EPI_ISL_687778\|2020-07 | ···················· | ···················· | ····················· | ···················· | ···················· | ···················· |
| V | 8320 hCoV-19/Japan/PG-6668/2020\|EPI_ISL_687779\|2020-07 | ···················· | ···················· | ····················· | ···················· | ···················· | ···················· |
| V | 8321 hCoV-19/Japan/PG-6669/2020\|EPI_ISL_687780\|2020-07 | ···················· | ···················· | ····················· | ···················· | ···················· | ···················· |
| V | 8322 hCoV-19/Japan/PG-6670/2020\|EPI_ISL_687781\|2020-07 | ···················· | ···················· | ····················· | ···················· | ···················· | ···················· |
| V | 8323 hCoV-19/Japan/PG-6671/2020\|EPI_ISL_687782\|2020-07 | ···················· | ···················· | ····················· | ···················· | ···················· | ···················· |
| V | 8324 hCoV-19/Japan/PG-6672/2020\|EPI_ISL_687783\|2020-07 | ···················· | ···················· | ····················· | ···················· | ···················· | ···················· |
| V | 8325 hCoV-19/Japan/PG-6673/2020\|EPI_ISL_687784\|2020-07 | ···················· | ···················· | ····················· | ···················· | ···················· | ···················· |
| V | 8326 hCoV-19/Japan/PG-6674/2020\|EPI_ISL_687785\|2020-07 | ···················· | ···················· | ····················· | ···················· | ···················· | ···················· |
| V | 8327 hCoV-19/Japan/PG-6675/2020\|EPI_ISL_687786\|2020-07 | ···················· | ···················· | ····················· | ···················· | ···················· | ···················· |
| V | 8328 hCoV-19/Japan/PG-6676/2020\|EPI_ISL_687787\|2020-07 | ···················· | ···················· | ····················· | ···················· | ···················· | ···················· |
| V | 8329 hCoV-19/Japan/PG-6677/2020\|EPI_ISL_687788\|2020-07 | ···················· | ···················· | ····················· | ···················· | ···················· | ···················· |
| V | 8330 hCoV-19/Japan/PG-6678/2020\|EPI_ISL_687789\|2020-07 | ···················· | ···················· | ····················· | ···················· | ···················· | ···················· |
| V | 8331 hCoV-19/Japan/PG-6679/2020\|EPI_ISL_687790\|2020-07 | ···················· | ···················· | ····················· | ···················· | ···················· | ···················· |
| V | 8332 hCoV-19/Japan/PG-6680/2020\|EPI_ISL_687791\|2020-07 | ···················· | ···················· | ····················· | ···················· | ···················· | ···················· |
| V | 8333 hCoV-19/Japan/PG-6681/2020\|EPI_ISL_687792\|2020-07 | ···················· | ···················· | ····················· | ···················· | ···················· | ···················· |
| V | 8334 hCoV-19/Japan/PG-6682/2020\|EPI_ISL_687793\|2020-07 | ···················· | ···················· | ····················· | ···················· | ···················· | ···················· |
| V | 8335 hCoV-19/Japan/PG-6683/2020\|EPI_ISL_687794\|2020-07 | ···················· | ···················· | ····················· | ···················· | ···················· | ···················· |
| V | 8336 hCoV-19/Japan/PG-6684/2020\|EPI_ISL_687795\|2020-07 | ···················· | ···················· | ····················· | ···················· | ···················· | ···················· |
| V | 8337 hCoV-19/Japan/PG-6685/2020\|EPI_ISL_687796\|2020-07 | ···················· | ···················· | ····················· | ···················· | ···················· | ···················· |
| V | 8338 hCoV-19/Japan/PG-6686/2020\|EPI_ISL_687797\|2020-07 | ···················· | ···················· | ····················· | ···················· | ···················· | ···················· |
| V | 8339 hCoV-19/Japan/PG-6687/2020\|EPI_ISL_687798\|2020-07 | ···················· | ···················· | ····················· | ···················· | ···················· | ···················· |
| V | 8340 hCoV-19/Japan/PG-6688/2020\|EPI_ISL_687799\|2020-07 | ···················· | ···················· | ····················· | ···················· | ···················· | ···················· |
| V | 8341 hCoV-19/Japan/PG-3962/2020\|EPI_ISL_692210\|2020-07 | ···················· | ···················· | ····················· | ···················· | ···················· | ···················· |
| V | 8342 hCoV-19/Japan/PG-3963/2020\|EPI_ISL_692211\|2020-07 | ···················· | ···················· | ····················· | ···················· | ···················· | ···················· |
| V | 8343 hCoV-19/Japan/PG-3964/2020\|EPI_ISL_692212\|2020-07 | ···················· | ···················· | ····················· | ···················· | ···················· | ···················· |
| V | 8344 hCoV-19/Japan/PG-3965/2020\|EPI_ISL_692213\|2020-07 | ···················· | ···················· | ····················· | ···················· | ···················· | ···················· |
| V | 8345 hCoV-19/Japan/PG-3966/2020\|EPI_ISL_692214\|2020-07 | ···················· | ···················· | ····················· | ···················· | ···················· | ···················· |
| V | 8346 hCoV-19/Japan/PG-3967/2020\|EPI_ISL_692215\|2020-07 | ···················· | ···················· | ····················· | ···················· | ···················· | ···················· |
| V | 8347 hCoV-19/Japan/PG-3968/2020\|EPI_ISL_692216\|2020-07 | ···················· | ···················· | ····················· | ···················· | ···················· | ···················· |
| V | 8348 hCoV-19/Japan/PG-3971/2020\|EPI_ISL_692217\|2020-07 | ···················· | ···················· | ····················· | ···················· | ···················· | ···················· |
| V | 8349 hCoV-19/Japan/PG-3976/2020\|EPI_ISL_692218\|2020-07 | ···················· | ···················· | ····················· | ···················· | ···················· | ···················· |
| V | 8350 hCoV-19/Japan/PG-3977/2020\|EPI_ISL_692219\|2020-07 | ···················· | ···················· | ····················· | ···················· | ···················· | ···················· |
| V | 8351 hCoV-19/Japan/PG-3978/2020\|EPI_ISL_692220\|2020-07 | ···················· | ···················· | ····················· | ···················· | ···················· | ···················· |
| V | 8352 hCoV-19/Japan/PG-3979/2020\|EPI_ISL_692221\|2020-07 | ···················· | ·A·················· | ····················· | ···················· | ···················· | ···················· |
| V | 8353 hCoV-19/Japan/PG-3980/2020\|EPI_ISL_692222\|2020-07 | ···················· | ···················· | ····················· | ···················· | ···················· | ···················· |
| V | 8354 hCoV-19/Japan/PG-3981/2020\|EPI_ISL_692223\|2020-07 | ···················· | ···················· | ····················· | ···················· | ···················· | ···················· |
| V | 8355 hCoV-19/Japan/PG-3982/2020\|EPI_ISL_692224\|2020-07 | ···················· | ···················· | ····················· | ···················· | ···················· | ···················· |
| V | 8356 hCoV-19/Japan/PG-3983/2020\|EPI_ISL_692225\|2020-07 | ···················· | ···················· | ····················· | ···················· | ···················· | ···················· |
| V | 8357 hCoV-19/Japan/PG-3984/2020\|EPI_ISL_692226\|2020-07 | ···················· | ···················· | ····················· | ···················· | ···················· | ···················· |
| V | 8358 hCoV-19/Japan/PG-3985/2020\|EPI_ISL_692227\|2020-07 | ···················· | ···················· | ····················· | ···················· | ···················· | ···················· |
| V | 8360 hCoV-19/Japan/PG-3988/2020\|EPI_ISL_692229\|2020-07 | ···················· | ·A·················· | ····················· | ···················· | ···················· | ···················· |
| V | 8361 hCoV-19/Japan/PG-3989/2020\|EPI_ISL_692230\|2020-07 | ···················· | ···················· | ····················· | ···················· | ···················· | ···················· |
| V | 8362 hCoV-19/Japan/PG-3990/2020\|EPI_ISL_692231\|2020-07 | ···················· | ···················· | ····················· | ···················· | ···················· | ···················· |
| V | 8363 hCoV-19/Japan/PG-4057/2020\|EPI_ISL_692298\|2020-07 | ···················· | ···················· | ····················· | ···················· | ···················· | ···················· |
| V | 8364 hCoV-19/Japan/PG-4058/2020\|EPI_ISL_692299\|2020-07 | ···················· | ···················· | ····················· | ···················· | ···················· | ···················· |
| V | 8365 hCoV-19/Japan/PG-4059/2020\|EPI_ISL_692300\|2020-07 | ···················· | ···················· | ····················· | ···················· | ···················· | ···················· |
| V | 8366 hCoV-19/Japan/PG-4060/2020\|EPI_ISL_692301\|2020-07 | ···················· | ···················· | ····················· | ···················· | ···················· | ···················· |
| V | 8367 hCoV-19/Japan/PG-4061/2020\|EPI_ISL_692302\|2020-07 | ···················· | ···················· | ····················· | ···················· | ···················· | ···················· |
| V | 8368 hCoV-19/Japan/PG-4062/2020\|EPI_ISL_692303\|2020-07 | ···················· | ···················· | ····················· | ···················· | ···················· | ···················· |
| V | 8369 hCoV-19/Japan/PG-4063/2020\|EPI_ISL_692304\|2020-07 | ···················· | ···················· | ····················· | ···················· | ···················· | ···················· |
| V | 8370 hCoV-19/Japan/PG-4064/2020\|EPI_ISL_692305\|2020-07 | ···················· | ···················· | ····················· | ···················· | ···················· | ···················· |
| V | 8371 hCoV-19/Japan/PG-4065/2020\|EPI_ISL_692306\|2020-07 | ···················· | ···················· | ····················· | ···················· | ···················· | ···················· |
| V | 8372 hCoV-19/Japan/PG-4066/2020\|EPI_ISL_692307\|2020-07 | ···················· | ···················· | ····················· | ···················· | ···················· | ···················· |
| V | 8373 hCoV-19/Japan/PG-4067/2020\|EPI_ISL_692308\|2020-07 | ···················· | ···················· | ····················· | ···················· | ···················· | ···················· |
| V | 8374 hCoV-19/Japan/PG-4068/2020\|EPI_ISL_692309\|2020-07 | ···················· | ···················· | ····················· | ···················· | ···················· | ···················· |
| V | 8375 hCoV-19/Japan/PG-4069/2020\|EPI_ISL_692310\|2020-07 | ···················· | ···················· | ····················· | ···················· | ···················· | ···················· |
| V | 8376 hCoV-19/Japan/PG-4070/2020\|EPI_ISL_692311\|2020-07 | ···················· | ···················· | ····················· | ···················· | ···················· | ···················· |
| V | 8377 hCoV-19/Japan/PG-4071/2020\|EPI_ISL_692312\|2020-07 | ···················· | ···················· | ····················· | ···················· | ···················· | ···················· |
| V | 8378 hCoV-19/Japan/PG-4072/2020\|EPI_ISL_692313\|2020-07 | ···················· | ···················· | ····················· | ···················· | ···················· | ···················· |
| V | 8379 hCoV-19/Japan/PG-4073/2020\|EPI_ISL_692314\|2020-07 | ···················· | ···················· | ····················· | ···················· | ···················· | ···················· |
| V | 8380 hCoV-19/Japan/PG-4074/2020\|EPI_ISL_692315\|2020-07 | ···················· | ···················· | ····················· | ···················· | ···················· | ···················· |
| V | 8381 hCoV-19/Japan/PG-4075/2020\|EPI_ISL_692316\|2020-07 | ···················· | ···················· | ····················· | ···················· | ···················· | ···················· |
| V | 8382 hCoV-19/Japan/PG-4076/2020\|EPI_ISL_692317\|2020-07 | ···················· | ···················· | ····················· | ···················· | ···················· | ···················· |
| V | 8383 hCoV-19/Japan/PG-4077/2020\|EPI_ISL_692318\|2020-07 | ···················· | ···················· | ····················· | ···················· | ···················· | ···················· |
| V | 8384 hCoV-19/Japan/PG-4078/2020\|EPI_ISL_692319\|2020-07 | ···················· | ···················· | ····················· | ···················· | ···················· | ···················· |
| V | 8385 hCoV-19/Japan/PG-4079/2020\|EPI_ISL_692320\|2020-07 | ···················· | ···················· | ····················· | ···················· | ···················· | ···················· |
| V | 8386 hCoV-19/Japan/PG-4080/2020\|EPI_ISL_692321\|2020-07 | ···················· | ···················· | ····················· | ···················· | ···················· | ···················· |
| V | 8387 hCoV-19/Japan/PG-4081/2020\|EPI_ISL_692322\|2020-07 | ···················· | ···················· | ····················· | ···················· | ···················· | ···················· |
| V | 8388 hCoV-19/Japan/PG-4082/2020\|EPI_ISL_692323\|2020-07 | ···················· | ···················· | ····················· | ···················· | ···················· | ···················· |
| V | 8389 hCoV-19/Japan/PG-4083/2020\|EPI_ISL_692324\|2020-07 | ···················· | ···················· | ····················· | ···················· | ···················· | ···················· |
| V | 8390 hCoV-19/Japan/PG-4084/2020\|EPI_ISL_692325\|2020-07 | ···················· | ···················· | ····················· | ···················· | ···················· | ···················· |
| V | 8391 hCoV-19/Japan/PG-4085/2020\|EPI_ISL_692326\|2020-07 | ···················· | ···················· | ····················· | ···················· | ···················· | ···················· |
| V | 8392 hCoV-19/Japan/PG-4086/2020\|EPI_ISL_692327\|2020-08 | ···················· | ···················· | ····················· | ···················· | ···················· | ···················· |
| V | 8393 hCoV-19/Japan/PG-4087/2020\|EPI_ISL_692328\|2020-07 | ···················· | ···················· | ····················· | ···················· | ···················· | ···················· |
| V | 8394 hCoV-19/Japan/PG-4088/2020\|EPI_ISL_692329\|2020-07 | ···················· | ···················· | ····················· | ···················· | ···················· | ···················· |
| V | 8395 hCoV-19/Japan/PG-4089/2020\|EPI_ISL_692330\|2020-07 | ···················· | ···················· | ····················· | ···················· | ···················· | ···················· |
| V | 8396 hCoV-19/Portugal/PT1586/2020\|EPI_ISL_693542\|2020-0 | ···················· | ···················· | ····················· | ···················· | ···················· | ···················· |

| | | | | | | | |
|---|---|---|---|---|---|---|---|
| V | 8397 hCoV-19/Portugal/PT1523/2020\|EPI_ISL_693543\|2020-0 | .................... | .................... | ..................... | .................... | .................... | .................... |
| V | 8398 hCoV-19/Portugal/PT1524/2020\|EPI_ISL_693544\|2020-0 | .................... | .................... | ..................... | .................... | .................... | .................... |
| V | 8399 hCoV-19/Portugal/PT1526/2020\|EPI_ISL_693545\|2020-0 | .................... | .................... | ..................... | .................... | .................... | .................... |
| V | 8400 hCoV-19/Portugal/PT1527/2020\|EPI_ISL_693546\|2020-0 | .................... | .................... | ..................... | .................... | .................... | .................... |
| V | 8401 hCoV-19/Portugal/PT1528/2020\|EPI_ISL_693547\|2020-0 | .................... | .................... | ..................... | .................... | .................... | .................... |
| V | 8402 hCoV-19/Portugal/PT1529/2020\|EPI_ISL_693548\|2020-0 | .................... | .................... | ..................... | .................... | .................... | .................... |
| V | 8403 hCoV-19/Portugal/PT1530/2020\|EPI_ISL_693549\|2020-0 | .................... | .................... | ..................... | .................... | .................... | .................... |
| V | 8404 hCoV-19/Portugal/PT1532/2020\|EPI_ISL_693550\|2020-0 | .................... | .................... | ..................... | .................... | .................... | .................... |
| V | 8405 hCoV-19/Portugal/PT1535/2020\|EPI_ISL_693551\|2020-0 | .................... | .................... | ..................... | .................... | .................... | .................... |
| V | 8406 hCoV-19/Portugal/PT1536/2020\|EPI_ISL_693552\|2020-0 | .................... | .................... | ..................... | .................... | .................... | .................... |
| V | 8407 hCoV-19/Portugal/PT1537/2020\|EPI_ISL_693553\|2020-0 | .................... | .................... | ..................... | .................... | .................... | .................... |
| V | 8408 hCoV-19/Portugal/PT1540/2020\|EPI_ISL_693554\|2020-0 | .................... | .................... | ..................... | .................... | .................... | .................... |
| V | 8409 hCoV-19/Portugal/PT1541/2020\|EPI_ISL_693555\|2020-0 | .................... | .................... | ..................... | .................... | .................... | .................... |
| V | 8410 hCoV-19/Portugal/PT1542/2020\|EPI_ISL_693556\|2020-0 | .................... | .................... | ..................... | .................... | .................... | .................... |
| V | 8411 hCoV-19/Portugal/PT1543/2020\|EPI_ISL_693557\|2020-0 | .................... | .................... | ..................... | .................... | .................... | .................... |
| V | 8412 hCoV-19/Portugal/PT1544/2020\|EPI_ISL_693558\|2020-0 | .................... | .................... | ..................... | .................... | .................... | .................... |
| V | 8413 hCoV-19/Portugal/PT1546/2020\|EPI_ISL_693559\|2020-0 | .................... | .................... | ..................... | .................... | .................... | .................... |
| V | 8414 hCoV-19/Portugal/PT1547/2020\|EPI_ISL_693560\|2020-0 | .................... | .................... | ..................... | .................... | .................... | .................... |
| V | 8415 hCoV-19/Portugal/PT1548/2020\|EPI_ISL_693561\|2020-0 | .................... | .................... | ..................... | .................... | .................... | .................... |
| V | 8416 hCoV-19/Portugal/PT1549/2020\|EPI_ISL_693562\|2020-0 | .................... | .................... | ..................... | .................... | .................... | .................... |
| V | 8417 hCoV-19/Portugal/PT1701/2020\|EPI_ISL_693630\|2020-0 | .................... | .........A.......... | ..................... | .................... | .................... | .................... |
| V | 8418 hCoV-19/Portugal/PT1702/2020\|EPI_ISL_693631\|2020-0 | .................... | .........A.......... | ..................... | .................... | .................... | .................... |
| V | 8419 hCoV-19/Portugal/PT1703/2020\|EPI_ISL_693632\|2020-0 | .................... | .........A.......... | ..................... | .................... | .................... | .................... |
| V | 8420 hCoV-19/Portugal/PT1704/2020\|EPI_ISL_693633\|2020-0 | .................... | .........A.......... | ..................... | .................... | .................... | .................... |
| V | 8421 hCoV-19/Portugal/PT1705/2020\|EPI_ISL_693634\|2020-0 | .................... | .........A.......... | ..................... | .................... | .................... | .................... |
| V | 8422 hCoV-19/Portugal/PT1706/2020\|EPI_ISL_693635\|2020-0 | .................... | .........A.......... | ..................... | .................... | .................... | .................... |
| V | 8423 hCoV-19/Portugal/PT1708/2020\|EPI_ISL_693636\|2020-0 | .................... | .........A.......... | ..................... | .................... | .................... | .................... |
| V | 8424 hCoV-19/Portugal/PT1710/2020\|EPI_ISL_693637\|2020-0 | .................... | .........A.......... | ..................... | .................... | .................... | .................... |
| V | 8425 hCoV-19/Portugal/PT1711/2020\|EPI_ISL_693638\|2020-0 | .................... | .........A.......... | ..................... | .................... | .................... | .................... |
| V | 8426 hCoV-19/Portugal/PT1712/2020\|EPI_ISL_693639\|2020-0 | .................... | .........A.......... | ..................... | .................... | .................... | .................... |
| V | 8427 hCoV-19/Portugal/PT1713/2020\|EPI_ISL_693640\|2020-0 | .................... | .........A.......... | ..................... | .................... | .................... | .................... |
| V | 8428 hCoV-19/Portugal/PT1714/2020\|EPI_ISL_693641\|2020-0 | .................... | .........A.......... | ..................... | .................... | .................... | .................... |
| V | 8429 hCoV-19/Portugal/PT1715/2020\|EPI_ISL_693642\|2020-0 | .................... | .........A.......... | ..................... | .................... | .................... | .................... |
| V | 8430 hCoV-19/Portugal/PT1716/2020\|EPI_ISL_693643\|2020-0 | .................... | .........A.......... | ..................... | .................... | .................... | .................... |
| V | 8431 hCoV-19/Portugal/PT1717/2020\|EPI_ISL_693644\|2020-0 | .................... | .........A.......... | ..................... | .................... | .................... | .................... |
| V | 8432 hCoV-19/Portugal/PT1718/2020\|EPI_ISL_693645\|2020-0 | .................... | .........A.......... | ..................... | .................... | .................... | .................... |
| V | 8433 hCoV-19/Portugal/PT1719/2020\|EPI_ISL_693646\|2020-0 | .................... | .........A.......... | ..................... | .................... | .................... | .................... |
| V | 8434 hCoV-19/Portugal/PT1720/2020\|EPI_ISL_693647\|2020-0 | .................... | .........A.......... | ..................... | .................... | .................... | .................... |
| V | 8435 hCoV-19/Portugal/PT1721/2020\|EPI_ISL_693648\|2020-0 | .................... | .........A.......... | ..................... | .................... | .................... | .................... |
| V | 8436 hCoV-19/Portugal/PT1722/2020\|EPI_ISL_693649\|2020-0 | .................... | .........A.......... | ..................... | .................... | .................... | .................... |
| V | 8437 hCoV-19/Portugal/PT1724/2020\|EPI_ISL_693650\|2020-0 | .................... | .........A.......... | ..................... | .................... | .................... | .................... |
| V | 8438 hCoV-19/Portugal/PT1725/2020\|EPI_ISL_693651\|2020-0 | .................... | .........A.......... | ..................... | .................... | .................... | .................... |
| V | 8439 hCoV-19/Portugal/PT1726/2020\|EPI_ISL_693652\|2020-0 | .................... | .........A.......... | ..................... | .................... | .................... | .................... |
| V | 8440 hCoV-19/Portugal/PT1727/2020\|EPI_ISL_693653\|2020-0 | .................... | .........A.......... | ..................... | .................... | .................... | .................... |
| V | 8441 hCoV-19/Portugal/PT1728/2020\|EPI_ISL_693654\|2020-0 | .................... | .........A.......... | ..................... | .................... | .................... | .................... |
| V | 8442 hCoV-19/Portugal/PT1731/2020\|EPI_ISL_693655\|2020-0 | .................... | .........A.......... | ..................... | .................... | .................... | .................... |
| V | 8443 hCoV-19/Portugal/PT1733/2020\|EPI_ISL_693656\|2020-0 | .................... | .........A.......... | ..................... | .................... | .................... | .................... |
| V | 8444 hCoV-19/Portugal/PT1736/2020\|EPI_ISL_693657\|2020-0 | .................... | .........A.......... | ..................... | .................... | .................... | .................... |
| V | 8445 hCoV-19/USA/MO-WUSTL-C1/2020\|EPI_ISL_693658\|2020-0 | .................... | .................... | ..................... | .................... | .................... | .................... |
| V | 8446 hCoV-19/USA/MO-WUSTL-C4/2020\|EPI_ISL_693659\|2020-0 | .................... | .................... | ..................... | .................... | .................... | .................... |
| V | 8447 hCoV-19/Czech Republic/NRL-10483/2020\|EPI_ISL_6936 | .................... | .................... | ..................... | .................... | .................... | .................... |
| V | 8448 hCoV-19/Czech Republic/NRL-10719/2020\|EPI_ISL_6936 | .................... | .................... | ..................... | .................... | .................... | .................... |
| V | 8449 hCoV-19/Japan/PG-1445/2020\|EPI_ISL_685052\|2020-04 | .................... | .................... | ..................... | .................... | .................... | .................... |
| V | 8450 hCoV-19/Japan/PG-1447/2020\|EPI_ISL_685054\|2020-04 | .................... | .................... | ..................... | .................... | .................... | .................... |
| V | 8451 hCoV-19/Japan/PG-1448/2020\|EPI_ISL_685055\|2020-04 | .................... | .................... | ..................... | .................... | .................... | .................... |
| V | 8452 hCoV-19/Japan/PG-1449/2020\|EPI_ISL_685056\|2020-04 | .................... | .................... | ..................... | .................... | .................... | .................... |
| V | 8453 hCoV-19/Japan/PG-1450/2020\|EPI_ISL_685057\|2020-04 | .................... | .................... | ..................... | .................... | .................... | .................... |
| V | 8454 hCoV-19/Japan/PG-1451/2020\|EPI_ISL_685058\|2020-04 | .................... | .................... | ..................... | .................... | .................... | .................... |
| V | 8455 hCoV-19/Japan/PG-1452/2020\|EPI_ISL_685059\|2020-04 | .................... | .................... | ..................... | .................... | .................... | .................... |
| V | 8456 hCoV-19/Japan/PG-1453/2020\|EPI_ISL_685060\|2020-04 | .................... | .................... | ..................... | .................... | .................... | .................... |
| V | 8457 hCoV-19/Japan/PG-1454/2020\|EPI_ISL_685061\|2020-04 | .................... | .................... | ..................... | .................... | .................... | .................... |
| V | 8458 hCoV-19/Japan/PG-1455/2020\|EPI_ISL_685062\|2020-04 | .................... | .................... | ..................... | .................... | .................... | .................... |
| V | 8459 hCoV-19/Japan/PG-1456/2020\|EPI_ISL_685063\|2020-04 | .................... | .................... | ..................... | .................... | .................... | .................... |
| V | 8460 hCoV-19/Japan/PG-1457/2020\|EPI_ISL_685064\|2020-04 | .................... | .................... | ..................... | .................... | .................... | .................... |
| V | 8461 hCoV-19/Japan/PG-1458/2020\|EPI_ISL_685065\|2020-04 | .................... | .................... | ..................... | .................... | .................... | .................... |
| V | 8463 hCoV-19/Japan/PG-1460/2020\|EPI_ISL_685067\|2020-04 | .................... | .................... | ..................... | .................... | .................... | .................... |
| V | 8464 hCoV-19/Japan/PG-1461/2020\|EPI_ISL_685068\|2020-04 | .................... | .................... | ..................... | .................... | .................... | .................... |
| V | 8465 hCoV-19/Japan/PG-1462/2020\|EPI_ISL_685069\|2020-04 | .................... | .................... | ..................... | .................... | .................... | .................... |
| V | 8466 hCoV-19/Japan/PG-1463/2020\|EPI_ISL_685070\|2020-04 | .................... | .................... | ..................... | .................... | .................... | .................... |
| V | 8467 hCoV-19/Japan/PG-1464/2020\|EPI_ISL_685071\|2020-04 | .................... | .................... | ..................... | .................... | .................... | .................... |
| V | 8468 hCoV-19/Japan/PG-1465/2020\|EPI_ISL_685072\|2020-04 | .................... | .................... | ..................... | .................... | .................... | .................... |
| V | 8469 hCoV-19/Japan/PG-1466/2020\|EPI_ISL_685073\|2020-04 | .................... | .................... | ..................... | .................... | .................... | .................... |
| V | 8470 hCoV-19/Japan/PG-1467/2020\|EPI_ISL_685074\|2020-04 | .................... | .................... | ..................... | .................... | .................... | .................... |
| V | 8471 hCoV-19/Japan/PG-1468/2020\|EPI_ISL_685075\|2020-04 | .................... | .................... | ..................... | .................... | .................... | .................... |
| V | 8472 hCoV-19/Japan/PG-1469/2020\|EPI_ISL_685076\|2020-04 | .................... | .................... | ..................... | .................... | .................... | .................... |
| V | 8473 hCoV-19/Japan/PG-1470/2020\|EPI_ISL_685077\|2020-04 | .................... | .................... | ..................... | .................... | .................... | .................... |
| V | 8474 hCoV-19/Japan/PG-1471/2020\|EPI_ISL_685078\|2020-04 | .................... | .................... | ..................... | .................... | .................... | .................... |
| V | 8475 hCoV-19/Japan/PG-1472/2020\|EPI_ISL_685079\|2020-04 | .................... | .................... | ..................... | .................... | .................... | .................... |
| V | 8476 hCoV-19/Japan/PG-1473/2020\|EPI_ISL_685080\|2020-04 | .................... | .................... | ..................... | .................... | .................... | .................... |
| V | 8477 hCoV-19/Japan/PG-1474/2020\|EPI_ISL_685081\|2020-04 | .................... | .................... | ..................... | .................... | .................... | .................... |

| | | | | | | | | |
|---|---|---|---|---|---|---|---|---|
| V | 8478 | hCoV-19/Japan/PG-1475/2020\|EPI_ISL_685082\|2020-04 | .................... | .................... | ..................... | .................... | .................... | .................... |
| V | 8479 | hCoV-19/Japan/PG-1476/2020\|EPI_ISL_685083\|2020-04 | .................... | .................... | ..................... | .................... | .................... | .................... |
| V | 8480 | hCoV-19/Japan/PG-1477/2020\|EPI_ISL_685084\|2020-04 | .................... | .................... | ..................... | .................... | .................... | .................... |
| V | 8481 | hCoV-19/Japan/PG-1478/2020\|EPI_ISL_685085\|2020-04 | .................... | .................... | ..................... | .................... | .................... | .................... |
| V | 8482 | hCoV-19/Japan/PG-1479/2020\|EPI_ISL_685086\|2020-04 | .................... | .................... | ..................... | .................... | .................... | .................... |
| V | 8483 | hCoV-19/Japan/PG-1480/2020\|EPI_ISL_685087\|2020-04 | .................... | .................... | ..................... | .................... | .................... | .................... |
| V | 8485 | hCoV-19/Japan/PG-1482/2020\|EPI_ISL_685089\|2020-04 | .................... | .................... | ..................... | .................... | .................... | .................... |
| V | 8486 | hCoV-19/Japan/PG-1493/2020\|EPI_ISL_685100\|2020-04 | .................... | .................... | ..................... | .................... | .................... | .................... |
| V | 8487 | hCoV-19/Japan/PG-1494/2020\|EPI_ISL_685101\|2020-04 | .................... | .................... | ..................... | .................... | .................... | .................... |
| V | 8488 | hCoV-19/Japan/PG-1496/2020\|EPI_ISL_685103\|2020-04 | .................... | .................... | ..................... | .................... | .................... | .................... |
| V | 8489 | hCoV-19/Japan/PG-8795/2020\|EPI_ISL_686383\|2020-08 | .................... | .................... | ..................... | .................... | .................... | .................... |
| V | 8490 | hCoV-19/Japan/PG-8797/2020\|EPI_ISL_686385\|2020-08 | .................... | .................... | ..................... | .................... | .................... | .................... |
| V | 8491 | hCoV-19/Japan/PG-8799/2020\|EPI_ISL_686386\|2020-08 | .................... | .................... | ..................... | .................... | .................... | .................... |
| V | 8492 | hCoV-19/Japan/PG-8800/2020\|EPI_ISL_686387\|2020-08 | .................... | .................... | ..................... | .................... | .................... | .................... |
| V | 8493 | hCoV-19/Japan/PG-8801/2020\|EPI_ISL_686388\|2020-08 | .................... | .................... | ..................... | .................... | .................... | .................... |
| V | 8494 | hCoV-19/Japan/PG-8802/2020\|EPI_ISL_686389\|2020-08 | .................... | .................... | ..................... | .................... | .................... | .................... |
| V | 8495 | hCoV-19/Japan/PG-8803/2020\|EPI_ISL_686390\|2020-08 | .................... | .................... | ..................... | .................... | .................... | .................... |
| V | 8496 | hCoV-19/Japan/PG-8804/2020\|EPI_ISL_686391\|2020-08 | .................... | .................... | ..................... | .................... | .................... | .................... |
| V | 8497 | hCoV-19/Japan/PG-8805/2020\|EPI_ISL_686392\|2020-08 | .................... | .................... | ..................... | .................... | .................... | .................... |
| V | 8498 | hCoV-19/Japan/PG-8807/2020\|EPI_ISL_686393\|2020-08 | .................... | .................... | ..................... | .................... | .................... | .................... |
| V | 8499 | hCoV-19/Japan/PG-8809/2020\|EPI_ISL_686394\|2020-08 | .................... | .................... | ..................... | .................... | .................... | .................... |
| V | 8500 | hCoV-19/Japan/PG-8810/2020\|EPI_ISL_686395\|2020-08 | .................... | .................... | ..................... | .................... | .................... | .................... |
| V | 8501 | hCoV-19/Japan/PG-8812/2020\|EPI_ISL_686396\|2020-08 | .................... | .................... | ..................... | .................... | .................... | .................... |
| V | 8502 | hCoV-19/Japan/PG-8815/2020\|EPI_ISL_686397\|2020-08 | .................... | .................... | ..................... | .................... | .................... | .................... |
| V | 8503 | hCoV-19/Japan/PG-8816/2020\|EPI_ISL_686398\|2020-08 | .................... | .................... | ..................... | .................... | .................... | .................... |
| V | 8504 | hCoV-19/Japan/PG-8817/2020\|EPI_ISL_686399\|2020-08 | .................... | .................... | ..................... | .................... | .................... | .................... |
| V | 8505 | hCoV-19/Japan/PG-8818/2020\|EPI_ISL_686400\|2020-08 | .................... | .................... | ..................... | .................... | .................... | .................... |
| V | 8506 | hCoV-19/Japan/PG-8819/2020\|EPI_ISL_686401\|2020-08 | .................... | .................... | ..................... | .................... | .................... | .................... |
| V | 8507 | hCoV-19/Japan/PG-8820/2020\|EPI_ISL_686402\|2020-08 | .................... | .................... | ..................... | .................... | .................... | .................... |
| V | 8508 | hCoV-19/Japan/PG-8821/2020\|EPI_ISL_686403\|2020-08 | .................... | .................... | ..................... | .................... | .................... | .................... |
| V | 8509 | hCoV-19/Japan/PG-8822/2020\|EPI_ISL_686404\|2020-08 | .................... | .................... | ..................... | .................... | .................... | .................... |
| V | 8510 | hCoV-19/Japan/PG-8823/2020\|EPI_ISL_686405\|2020-08 | .................... | .................... | ..................... | .................... | .................... | .................... |
| V | 8511 | hCoV-19/Japan/PG-8824/2020\|EPI_ISL_686406\|2020-08 | .................... | .................... | ..................... | .................... | .................... | .................... |
| V | 8512 | hCoV-19/Japan/PG-8826/2020\|EPI_ISL_686407\|2020-08 | .................... | .................... | ..................... | .................... | .................... | .................... |
| V | 8513 | hCoV-19/Japan/PG-8827/2020\|EPI_ISL_686408\|2020-08 | .................... | .................... | ..................... | .................... | .................... | .................... |
| V | 8514 | hCoV-19/Japan/PG-8830/2020\|EPI_ISL_686409\|2020-09 | .................... | .................... | ..................... | .................... | .................... | .................... |
| V | 8515 | hCoV-19/Japan/PG-8832/2020\|EPI_ISL_686410\|2020-08 | .................... | .................... | ..................... | .................... | .................... | .................... |
| V | 8516 | hCoV-19/Japan/PG-8833/2020\|EPI_ISL_686411\|2020-08 | .................... | .................... | ..................... | .................... | .................... | .................... |
| V | 8517 | hCoV-19/Japan/PG-8834/2020\|EPI_ISL_686412\|2020-08 | .................... | .................... | ..................... | .................... | .................... | .................... |
| V | 8518 | hCoV-19/Japan/PG-8835/2020\|EPI_ISL_686413\|2020-08 | .................... | .................... | ..................... | .................... | .................... | .................... |
| V | 8519 | hCoV-19/Japan/PG-8836/2020\|EPI_ISL_686414\|2020-08 | .................... | .................... | ..................... | .................... | .................... | .................... |
| V | 8520 | hCoV-19/Japan/PG-8837/2020\|EPI_ISL_686415\|2020-08 | .................... | .................... | ..................... | .................... | .................... | .................... |
| V | 8521 | hCoV-19/Japan/PG-8838/2020\|EPI_ISL_686416\|2020-09 | .................... | .................... | ..................... | .................... | .................... | .................... |
| V | 8522 | hCoV-19/Japan/PG-8839/2020\|EPI_ISL_686417\|2020-09 | .................... | .................... | ..................... | .................... | .................... | .................... |
| V | 8523 | hCoV-19/Japan/PG-8872/2020\|EPI_ISL_686418\|2020-08 | .................... | .................... | ..................... | .................... | .................... | .................... |
| V | 8524 | hCoV-19/Japan/PG-8873/2020\|EPI_ISL_686419\|2020-08 | .................... | .................... | ..................... | .................... | .................... | .................... |
| V | 8525 | hCoV-19/Japan/PG-8874/2020\|EPI_ISL_686420\|2020-08 | .................... | .................... | ..................... | .................... | .................... | .................... |
| V | 8526 | hCoV-19/Japan/PG-8875/2020\|EPI_ISL_686421\|2020-08 | .................... | .................... | ..................... | .................... | .................... | .................... |
| V | 8527 | hCoV-19/Japan/PG-8876/2020\|EPI_ISL_686422\|2020-08 | .................... | .................... | ..................... | .................... | .................... | .................... |
| V | 8528 | hCoV-19/Japan/PG-8877/2020\|EPI_ISL_686423\|2020-08 | .................... | .................... | ..................... | .................... | .................... | .................... |
| V | 8529 | hCoV-19/Japan/PG-8878/2020\|EPI_ISL_686424\|2020-08 | .................... | .................... | ..................... | .................... | .................... | .................... |
| V | 8530 | hCoV-19/Japan/PG-8879/2020\|EPI_ISL_686425\|2020-08 | .................... | .................... | ..................... | .................... | .................... | .................... |
| V | 8531 | hCoV-19/Japan/PG-8880/2020\|EPI_ISL_686426\|2020-08 | .................... | .................... | ..................... | .................... | .................... | .................... |
| V | 8532 | hCoV-19/Japan/PG-8881/2020\|EPI_ISL_686427\|2020-08 | .................... | .................... | ..................... | .................... | .................... | .................... |
| V | 8533 | hCoV-19/Japan/PG-8883/2020\|EPI_ISL_686429\|2020-08 | .................... | .................... | ..................... | .................... | .................... | .................... |
| V | 8534 | hCoV-19/Japan/PG-8884/2020\|EPI_ISL_686430\|2020-08 | .................... | .................... | ..................... | .................... | .................... | .................... |
| V | 8535 | hCoV-19/Japan/PG-8885/2020\|EPI_ISL_686431\|2020-08 | .................... | .................... | ..................... | .................... | .................... | .................... |
| V | 8536 | hCoV-19/Japan/PG-8886/2020\|EPI_ISL_686432\|2020-08 | .................... | .................... | ..................... | .................... | .................... | .................... |
| V | 8537 | hCoV-19/Japan/PG-8888/2020\|EPI_ISL_686434\|2020-08 | .................... | .................... | ..................... | .................... | .................... | .................... |
| V | 8538 | hCoV-19/Japan/PG-6588/2020\|EPI_ISL_687700\|2020-07 | .................... | .................... | ..................... | .................... | .................... | .................... |
| V | 8539 | hCoV-19/Japan/PG-6589/2020\|EPI_ISL_687701\|2020-07 | .................... | .................... | ..................... | .................... | .................... | .................... |
| V | 8540 | hCoV-19/Japan/PG-6590/2020\|EPI_ISL_687702\|2020-07 | .................... | .................... | ..................... | .................... | .................... | .................... |
| V | 8541 | hCoV-19/Japan/PG-6591/2020\|EPI_ISL_687703\|2020-07 | .................... | .................... | ..................... | .................... | .................... | .................... |
| V | 8542 | hCoV-19/Japan/PG-6592/2020\|EPI_ISL_687704\|2020-07 | .................... | .................... | ..................... | .................... | .................... | .................... |
| V | 8543 | hCoV-19/Japan/PG-6593/2020\|EPI_ISL_687705\|2020-08 | .................... | .................... | ..................... | .................... | .................... | .................... |
| V | 8544 | hCoV-19/Japan/PG-6594/2020\|EPI_ISL_687706\|2020-08 | .................... | .................... | ..................... | .................... | .................... | .................... |
| V | 8545 | hCoV-19/Japan/PG-6595/2020\|EPI_ISL_687707\|2020-08 | .................... | .................... | ..................... | .................... | .................... | .................... |
| V | 8546 | hCoV-19/Japan/PG-6596/2020\|EPI_ISL_687708\|2020-08 | .................... | .................... | ..................... | .................... | .................... | .................... |
| V | 8547 | hCoV-19/Japan/PG-6597/2020\|EPI_ISL_687709\|2020-08 | .................... | .................... | ..................... | .................... | .................... | .................... |
| V | 8548 | hCoV-19/Japan/PG-6598/2020\|EPI_ISL_687710\|2020-08 | .................... | .................... | ..................... | .................... | .................... | .................... |
| V | 8549 | hCoV-19/Japan/PG-6599/2020\|EPI_ISL_687711\|2020-08 | .................... | .................... | ..................... | .................... | .................... | .................... |
| V | 8550 | hCoV-19/Japan/PG-6600/2020\|EPI_ISL_687712\|2020-08 | .................... | .................... | ..................... | .................... | .................... | .................... |
| V | 8551 | hCoV-19/Japan/PG-6601/2020\|EPI_ISL_687713\|2020-08 | .................... | .................... | ..................... | .................... | .................... | .................... |
| V | 8552 | hCoV-19/Japan/PG-6602/2020\|EPI_ISL_687714\|2020-08 | .................... | .................... | ..................... | .................... | .................... | .................... |
| V | 8553 | hCoV-19/Japan/PG-6603/2020\|EPI_ISL_687715\|2020-08 | .................... | .................... | ..................... | .................... | .................... | .................... |
| V | 8554 | hCoV-19/Japan/PG-6604/2020\|EPI_ISL_687716\|2020-08 | .................... | .................... | ..................... | .................... | .................... | .................... |
| V | 8555 | hCoV-19/Japan/PG-6605/2020\|EPI_ISL_687717\|2020-08 | .................... | .................... | ..................... | .................... | .................... | .................... |
| V | 8556 | hCoV-19/Japan/PG-6606/2020\|EPI_ISL_687718\|2020-06 | .................... | .................... | ..................... | .................... | .................... | .................... |
| V | 8557 | hCoV-19/Japan/PG-6607/2020\|EPI_ISL_687719\|2020-07 | .................... | .................... | ..................... | .................... | .................... | .................... |
| V | 8558 | hCoV-19/Japan/PG-6608/2020\|EPI_ISL_687720\|2020-07 | .................... | .................... | ..................... | .................... | .................... | .................... |

| | | | | | | | |
|---|---|---|---|---|---|---|---|
| V | 8559 hCoV-19/Japan/PG-6609/2020\|EPI_ISL_687721\|2020-07 | .................... | .................... | ..................... | .................... | .................... | .................... |
| V | 8560 hCoV-19/Japan/PG-6610/2020\|EPI_ISL_687722\|2020-07 | .................... | .................... | ..................... | .................... | .................... | .................... |
| V | 8561 hCoV-19/Japan/PG-6611/2020\|EPI_ISL_687723\|2020-07 | .................... | .................... | ..................... | .................... | .................... | .................... |
| V | 8562 hCoV-19/Japan/PG-6612/2020\|EPI_ISL_687724\|2020-07 | .................... | .................... | ..................... | .................... | .................... | .................... |
| V | 8563 hCoV-19/Japan/PG-6613/2020\|EPI_ISL_687725\|2020-07 | .................... | .................... | ..................... | .................... | .................... | .................... |
| V | 8564 hCoV-19/Japan/PG-6614/2020\|EPI_ISL_687726\|2020-07 | .................... | .................... | ..................... | .................... | .................... | .................... |
| V | 8565 hCoV-19/Japan/PG-6615/2020\|EPI_ISL_687727\|2020-07 | .................... | .................... | ..................... | .................... | .................... | .................... |
| V | 8566 hCoV-19/Japan/PG-6616/2020\|EPI_ISL_687728\|2020-07 | .................... | .................... | ..................... | .................... | .................... | .................... |
| V | 8567 hCoV-19/Japan/PG-6617/2020\|EPI_ISL_687729\|2020-07 | .................... | .................... | ..................... | .................... | .................... | .................... |
| V | 8568 hCoV-19/Japan/PG-6618/2020\|EPI_ISL_687730\|2020-07 | .................... | .................... | ..................... | .................... | .................... | .................... |
| V | 8569 hCoV-19/Japan/PG-6619/2020\|EPI_ISL_687731\|2020-07 | .................... | .................... | ..................... | .................... | .................... | .................... |
| V | 8570 hCoV-19/Japan/PG-6620/2020\|EPI_ISL_687732\|2020-07 | .................... | .................... | ..................... | .................... | .................... | .................... |
| V | 8571 hCoV-19/Japan/PG-6621/2020\|EPI_ISL_687733\|2020-08 | .................... | .................... | ..................... | .................... | .................... | .................... |
| V | 8572 hCoV-19/Japan/PG-6622/2020\|EPI_ISL_687734\|2020-08 | .................... | .................... | ..................... | .................... | .................... | .................... |
| V | 8573 hCoV-19/Japan/PG-6623/2020\|EPI_ISL_687735\|2020-08 | .................... | .................... | ..................... | .................... | .................... | .................... |
| V | 8574 hCoV-19/Japan/PG-6624/2020\|EPI_ISL_687736\|2020-08 | .................... | .................... | ..................... | .................... | .................... | .................... |
| V | 8575 hCoV-19/Japan/PG-6625/2020\|EPI_ISL_687737\|2020-08 | .................... | .................... | ..................... | .................... | .................... | .................... |
| V | 8576 hCoV-19/Japan/PG-6626/2020\|EPI_ISL_687738\|2020-08 | .................... | .................... | ..................... | .................... | .................... | .................... |
| V | 8577 hCoV-19/Japan/PG-6627/2020\|EPI_ISL_687739\|2020-08 | .................... | .................... | ..................... | .................... | .................... | .................... |
| V | 8578 hCoV-19/Japan/PG-6628/2020\|EPI_ISL_687740\|2020-08 | .................... | .................... | ..................... | .................... | .................... | .................... |
| V | 8579 hCoV-19/Japan/PG-6629/2020\|EPI_ISL_687741\|2020-08 | .................... | .................... | ..................... | .................... | .................... | .................... |
| V | 8580 hCoV-19/Japan/PG-6630/2020\|EPI_ISL_687742\|2020-08 | .................... | .................... | ..................... | .................... | .................... | .................... |
| V | 8581 hCoV-19/Japan/PG-6631/2020\|EPI_ISL_687743\|2020-09 | .................... | .................... | ..................... | .................... | .................... | .................... |
| V | 8582 hCoV-19/Japan/PG-6632/2020\|EPI_ISL_687744\|2020-09 | .................... | .................... | ..................... | .................... | .................... | .................... |
| V | 8583 hCoV-19/Japan/PG-6633/2020\|EPI_ISL_687745\|2020-09 | .................... | .................... | ..................... | .................... | .................... | .................... |
| V | 8584 hCoV-19/Japan/PG-6634/2020\|EPI_ISL_687746\|2020-06 | .................... | .................... | ..................... | .................... | .................... | .................... |
| V | 8585 hCoV-19/Japan/PG-6635/2020\|EPI_ISL_687747\|2020-06 | .................... | .................... | ..................... | .................... | .................... | .................... |
| V | 8586 hCoV-19/Japan/PG-6636/2020\|EPI_ISL_687748\|2020-06 | .................... | .................... | ..................... | .................... | .................... | .................... |
| V | 8587 hCoV-19/Japan/PG-6637/2020\|EPI_ISL_687749\|2020-06 | .................... | .................... | ..................... | .................... | .................... | .................... |
| V | 8588 hCoV-19/Japan/PG-6638/2020\|EPI_ISL_687750\|2020-06 | .................... | .................... | ..................... | .................... | .................... | .................... |
| V | 8589 hCoV-19/Japan/PG-6639/2020\|EPI_ISL_687751\|2020-06 | .................... | .................... | ..................... | .................... | .................... | .................... |
| V | 8590 hCoV-19/Japan/PG-6640/2020\|EPI_ISL_687752\|2020-06 | .................... | .................... | ..................... | .................... | .................... | .................... |
| V | 8591 hCoV-19/Japan/PG-6641/2020\|EPI_ISL_687753\|2020-06 | .................... | .................... | ..................... | .................... | .................... | .................... |
| V | 8592 hCoV-19/Japan/PG-6642/2020\|EPI_ISL_687754\|2020-07 | .................... | .................... | ..................... | .................... | .................... | .................... |
| V | 8593 hCoV-19/Japan/PG-6643/2020\|EPI_ISL_687755\|2020-07 | .................... | .................... | ..................... | .................... | .................... | .................... |
| V | 8594 hCoV-19/Japan/PG-6644/2020\|EPI_ISL_687756\|2020-07 | .................... | .................... | ..................... | .................... | .................... | .................... |
| V | 8595 hCoV-19/Japan/PG-6645/2020\|EPI_ISL_687757\|2020-07 | .................... | .................... | ..................... | .................... | .................... | .................... |
| V | 8596 hCoV-19/Japan/PG-6646/2020\|EPI_ISL_687758\|2020-07 | .................... | .................... | ..................... | .................... | .................... | .................... |
| V | 8597 hCoV-19/Japan/PG-6647/2020\|EPI_ISL_687759\|2020-07 | .................... | .................... | ..................... | .................... | .................... | .................... |
| V | 8598 hCoV-19/Japan/PG-6648/2020\|EPI_ISL_687760\|2020-07 | .................... | .................... | ..................... | .................... | .................... | .................... |
| V | 8599 hCoV-19/Japan/PG-6649/2020\|EPI_ISL_687761\|2020-07 | .................... | .................... | ..................... | .................... | .................... | .................... |
| V | 8600 hCoV-19/Japan/PG-6650/2020\|EPI_ISL_687762\|2020-07 | .................... | .................... | ..................... | .................... | .................... | .................... |
| V | 8601 hCoV-19/Japan/PG-6651/2020\|EPI_ISL_687763\|2020-07 | .................... | .................... | ..................... | .................... | .................... | .................... |
| V | 8602 hCoV-19/Japan/PG-3991/2020\|EPI_ISL_692232\|2020-07 | .................... | .................... | ..................... | .................... | .................... | .................... |
| V | 8603 hCoV-19/Japan/PG-3992/2020\|EPI_ISL_692233\|2020-07 | .................... | .................... | ..................... | .................... | .................... | .................... |
| V | 8604 hCoV-19/Japan/PG-3993/2020\|EPI_ISL_692234\|2020-07 | .................... | .................... | ..................... | .................... | .................... | .................... |
| V | 8605 hCoV-19/Japan/PG-3994/2020\|EPI_ISL_692235\|2020-07 | .................... | .................... | ..................... | .................... | .................... | .................... |
| V | 8606 hCoV-19/Japan/PG-3995/2020\|EPI_ISL_692236\|2020-07 | .................... | .................... | ..................... | .................... | .................... | .................... |
| V | 8607 hCoV-19/Japan/PG-3996/2020\|EPI_ISL_692237\|2020-07 | .................... | .................... | ..................... | .................... | .................... | .................... |
| V | 8608 hCoV-19/Japan/PG-3997/2020\|EPI_ISL_692238\|2020-07 | .................... | .................... | ..................... | .................... | .................... | .................... |
| V | 8609 hCoV-19/Japan/PG-3998/2020\|EPI_ISL_692239\|2020-07 | .................... | .................... | ..................... | .................... | .................... | .................... |
| V | 8610 hCoV-19/Japan/PG-3999/2020\|EPI_ISL_692240\|2020-07 | .................... | .................... | ..................... | .................... | .................... | .................... |
| V | 8611 hCoV-19/Japan/PG-4000/2020\|EPI_ISL_692241\|2020-07 | .................... | .................... | ..................... | .................... | .................... | .................... |
| V | 8612 hCoV-19/Japan/PG-4001/2020\|EPI_ISL_692242\|2020-07 | .................... | .................... | ..................... | .................... | .................... | .................... |
| V | 8613 hCoV-19/Japan/PG-4002/2020\|EPI_ISL_692243\|2020-07 | .................... | .................... | ..................... | .................... | .................... | .................... |
| V | 8614 hCoV-19/Japan/PG-4003/2020\|EPI_ISL_692244\|2020-07 | .................... | .................... | ..................... | .................... | .................... | .................... |
| V | 8615 hCoV-19/Japan/PG-4004/2020\|EPI_ISL_692245\|2020-07 | .................... | .................... | ..................... | .................... | .................... | .................... |
| V | 8616 hCoV-19/Japan/PG-4005/2020\|EPI_ISL_692246\|2020-07 | .................... | .................... | ..................... | .................... | .................... | .................... |
| V | 8617 hCoV-19/Japan/PG-4006/2020\|EPI_ISL_692247\|2020-07 | .................... | .................... | ..................... | .................... | .................... | .................... |
| V | 8618 hCoV-19/Japan/PG-4007/2020\|EPI_ISL_692248\|2020-07 | .................... | .................... | ..................... | .................... | .................... | .................... |
| V | 8619 hCoV-19/Japan/PG-4008/2020\|EPI_ISL_692249\|2020-07 | .................... | .................... | ..................... | .................... | .................... | .................... |
| V | 8620 hCoV-19/Japan/PG-4009/2020\|EPI_ISL_692250\|2020-07 | .................... | .................... | ..................... | .................... | .................... | .................... |
| V | 8621 hCoV-19/Japan/PG-4010/2020\|EPI_ISL_692251\|2020-07 | .................... | .................... | ..................... | .................... | .................... | .................... |
| V | 8622 hCoV-19/Japan/PG-4011/2020\|EPI_ISL_692252\|2020-07 | .................... | .................... | ..................... | .................... | .................... | .................... |
| V | 8623 hCoV-19/Japan/PG-4012/2020\|EPI_ISL_692253\|2020-07 | .................... | .................... | ..................... | .................... | .................... | .................... |
| V | 8624 hCoV-19/Japan/PG-4013/2020\|EPI_ISL_692254\|2020-07 | .................... | .................... | ..................... | .................... | .................... | .................... |
| V | 8625 hCoV-19/Japan/PG-4014/2020\|EPI_ISL_692255\|2020-07 | .................... | .................... | ..................... | .................... | .................... | .................... |
| V | 8626 hCoV-19/Japan/PG-4015/2020\|EPI_ISL_692256\|2020-07 | .................... | .................... | ..................... | .................... | .................... | .................... |
| V | 8627 hCoV-19/Japan/PG-4016/2020\|EPI_ISL_692257\|2020-07 | .................... | .................... | ..................... | .................... | .................... | .................... |
| V | 8628 hCoV-19/Japan/PG-4017/2020\|EPI_ISL_692258\|2020-07 | .................... | .................... | ..................... | .................... | .................... | .................... |
| V | 8629 hCoV-19/Japan/PG-4018/2020\|EPI_ISL_692259\|2020-07 | .................... | .................... | ..................... | .................... | .................... | .................... |
| V | 8630 hCoV-19/Japan/PG-4019/2020\|EPI_ISL_692260\|2020-07 | .................... | .................... | ..................... | .................... | .................... | .................... |
| V | 8631 hCoV-19/Japan/PG-4020/2020\|EPI_ISL_692261\|2020-07 | .................... | .................... | ..................... | .................... | .................... | .................... |
| V | 8632 hCoV-19/Japan/PG-4021/2020\|EPI_ISL_692262\|2020-07 | .................... | .................... | ..................... | .................... | .................... | .................... |
| V | 8633 hCoV-19/Japan/PG-4022/2020\|EPI_ISL_692263\|2020-07 | .................... | .................... | ..................... | .................... | .................... | .................... |
| V | 8634 hCoV-19/Japan/PG-4023/2020\|EPI_ISL_692264\|2020-07 | .................... | .................... | ..................... | .................... | .................... | .................... |
| V | 8635 hCoV-19/Japan/PG-4024/2020\|EPI_ISL_692265\|2020-07 | .................... | .................... | ..................... | .................... | .................... | .................... |
| V | 8636 hCoV-19/Japan/PG-4025/2020\|EPI_ISL_692266\|2020-07 | .................... | .................... | ..................... | .................... | .................... | .................... |
| V | 8637 hCoV-19/Japan/PG-4026/2020\|EPI_ISL_692267\|2020-07 | .................... | .................... | ..................... | .................... | .................... | .................... |
| V | 8638 hCoV-19/Japan/PG-4027/2020\|EPI_ISL_692268\|2020-07 | .................... | .................... | ..................... | .................... | .................... | .................... |

| | | | | | | | | |
|---|---|---|---|---|---|---|---|---|
| V | 8639 hCoV-19/Japan/PG-4028/2020\|EPI_ISL_692269\|2020-07 | ···················· | ···················· | ···················· | ···················· | ···················· | ···················· | |
| V | 8640 hCoV-19/Japan/PG-4029/2020\|EPI_ISL_692270\|2020-07 | ···················· | ···················· | ···················· | ···················· | ···················· | ···················· | |
| V | 8641 hCoV-19/Japan/PG-4030/2020\|EPI_ISL_692271\|2020-07 | ···················· | ···················· | ···················· | ···················· | ···················· | ···················· | |
| V | 8642 hCoV-19/Japan/PG-4031/2020\|EPI_ISL_692272\|2020-07 | ···················· | ···················· | ···················· | ···················· | ···················· | ···················· | |
| V | 8643 hCoV-19/Japan/PG-4032/2020\|EPI_ISL_692273\|2020-07 | ···················· | ···················· | ···················· | ···················· | ···················· | ···················· | |
| V | 8644 hCoV-19/Japan/PG-4033/2020\|EPI_ISL_692274\|2020-07 | ···················· | ···················· | ···················· | ···················· | ···················· | ···················· | |
| V | 8645 hCoV-19/Japan/PG-4034/2020\|EPI_ISL_692275\|2020-07 | ···················· | ···················· | ···················· | ···················· | ···················· | ···················· | |
| V | 8646 hCoV-19/Japan/PG-4035/2020\|EPI_ISL_692276\|2020-07 | ···················· | ···················· | ···················· | ···················· | ···················· | ···················· | |
| V | 8647 hCoV-19/Japan/PG-4036/2020\|EPI_ISL_692277\|2020-07 | ···················· | ···················· | ···················· | ···················· | ···················· | ···················· | |
| V | 8648 hCoV-19/Japan/PG-4037/2020\|EPI_ISL_692278\|2020-07 | ···················· | ···················· | ···················· | ···················· | ···················· | ···················· | |
| V | 8649 hCoV-19/Japan/PG-4038/2020\|EPI_ISL_692279\|2020-07 | ···················· | ···················· | ···················· | ···················· | ···················· | ···················· | |
| V | 8650 hCoV-19/Japan/PG-4039/2020\|EPI_ISL_692280\|2020-07 | ···················· | ···················· | ···················· | ···················· | ···················· | ···················· | |
| V | 8651 hCoV-19/Japan/PG-4040/2020\|EPI_ISL_692281\|2020-07 | ···················· | ···················· | ···················· | ···················· | ···················· | ···················· | |
| V | 8652 hCoV-19/Japan/PG-4041/2020\|EPI_ISL_692282\|2020-07 | ···················· | ···················· | ···················· | ···················· | ···················· | ···················· | |
| V | 8653 hCoV-19/Japan/PG-4042/2020\|EPI_ISL_692283\|2020-07 | ···················· | ···················· | ···················· | ···················· | ···················· | ···················· | |
| V | 8654 hCoV-19/Japan/PG-4043/2020\|EPI_ISL_692284\|2020-07 | ···················· | ···················· | ···················· | ···················· | ···················· | ···················· | |
| V | 8655 hCoV-19/Japan/PG-4044/2020\|EPI_ISL_692285\|2020-07 | ···················· | ···················· | ···················· | ···················· | ···················· | ···················· | |
| V | 8656 hCoV-19/Japan/PG-4045/2020\|EPI_ISL_692286\|2020-07 | ···················· | ···················· | ···················· | ···················· | ···················· | ···················· | |
| V | 8657 hCoV-19/Japan/PG-4046/2020\|EPI_ISL_692287\|2020-07 | ···················· | ···················· | ···················· | ···················· | ···················· | ···················· | |
| V | 8658 hCoV-19/Japan/PG-4047/2020\|EPI_ISL_692288\|2020-07 | ···················· | ···················· | ···················· | ···················· | ···················· | ···················· | |
| V | 8659 hCoV-19/Japan/PG-4048/2020\|EPI_ISL_692289\|2020-07 | ···················· | ···················· | ···················· | ···················· | ···················· | ···················· | |
| V | 8660 hCoV-19/Japan/PG-4049/2020\|EPI_ISL_692290\|2020-07 | ···················· | ···················· | ···················· | ···················· | ···················· | ···················· | |
| V | 8661 hCoV-19/Japan/PG-4050/2020\|EPI_ISL_692291\|2020-07 | ···················· | ···················· | ···················· | ···················· | ···················· | ···················· | |
| V | 8662 hCoV-19/Japan/PG-4051/2020\|EPI_ISL_692292\|2020-07 | ···················· | ···················· | ···················· | ···················· | ···················· | ···················· | |
| V | 8663 hCoV-19/Japan/PG-4052/2020\|EPI_ISL_692293\|2020-07 | ···················· | ···················· | ···················· | ···················· | ···················· | ···················· | |
| V | 8664 hCoV-19/Japan/PG-4053/2020\|EPI_ISL_692294\|2020-07 | ···················· | ···················· | ···················· | ···················· | ···················· | ···················· | |
| V | 8665 hCoV-19/Japan/PG-4054/2020\|EPI_ISL_692295\|2020-07 | ···················· | ···················· | ···················· | ···················· | ···················· | ···················· | |
| V | 8666 hCoV-19/Japan/PG-4055/2020\|EPI_ISL_692296\|2020-07 | ···················· | ···················· | ···················· | ···················· | ···················· | ···················· | |
| V | 8667 hCoV-19/Japan/PG-4056/2020\|EPI_ISL_692297\|2020-07 | ···················· | ···················· | ···················· | ···················· | ···················· | ···················· | |
| V | 8670 hCoV-19/Portugal/PT1552/2020\|EPI_ISL_693565\|2020-0 | ···················· | ···················· | ···················· | ···················· | ···················· | ···················· | |
| V | 8671 hCoV-19/Portugal/PT1553/2020\|EPI_ISL_693566\|2020-0 | ···················· | ···················· | ···················· | ···················· | ···················· | ···················· | |
| V | 8672 hCoV-19/Portugal/PT1554/2020\|EPI_ISL_693567\|2020-0 | ···················· | ···················· | ···················· | ···················· | ···················· | ···················· | |
| V | 8673 hCoV-19/Portugal/PT1555/2020\|EPI_ISL_693568\|2020-0 | ···················· | ···················· | ···················· | ···················· | ···················· | ···················· | |
| V | 8674 hCoV-19/Portugal/PT1558/2020\|EPI_ISL_693569\|2020-0 | ···················· | ···················· | ···················· | ···················· | ···················· | ···················· | |
| V | 8675 hCoV-19/Portugal/PT1559/2020\|EPI_ISL_693570\|2020-0 | ···················· | ···················· | ···················· | ···················· | ···················· | ···················· | |
| V | 8676 hCoV-19/Portugal/PT1560/2020\|EPI_ISL_693571\|2020-0 | ···················· | ···················· | ···················· | ···················· | ···················· | ···················· | |
| V | 8677 hCoV-19/Portugal/PT1561/2020\|EPI_ISL_693572\|2020-0 | ···················· | ···················· | ···················· | ···················· | ···········N········ | ···················· | |
| V | 8678 hCoV-19/Portugal/PT1562/2020\|EPI_ISL_693573\|2020-0 | ···················· | ···················· | ···················· | ···················· | ···················· | ···················· | |
| V | 8679 hCoV-19/Portugal/PT1564/2020\|EPI_ISL_693574\|2020-0 | ···················· | ···················· | ···················· | ···················· | ···················· | ···················· | |
| V | 8680 hCoV-19/Portugal/PT1565/2020\|EPI_ISL_693575\|2020-0 | ···················· | ···················· | ···················· | ···················· | ···················· | ···················· | |
| V | 8681 hCoV-19/Portugal/PT1566/2020\|EPI_ISL_693576\|2020-0 | ···················· | ···················· | ···················· | ···················· | ···················· | ···················· | |
| V | 8682 hCoV-19/Portugal/PT1567/2020\|EPI_ISL_693577\|2020-0 | ···················· | ···················· | ···················· | ···················· | ···················· | ···················· | |
| V | 8684 hCoV-19/Portugal/PT1571/2020\|EPI_ISL_693579\|2020-0 | ···················· | ···················· | ···················· | ···················· | ···················· | ···················· | |
| V | 8685 hCoV-19/Portugal/PT1572/2020\|EPI_ISL_693580\|2020-0 | ···················· | ···················· | ···················· | ···················· | ···················· | ···················· | |
| V | 8686 hCoV-19/Portugal/PT1573/2020\|EPI_ISL_693581\|2020-0 | ···················· | ···················· | ···················· | ···················· | ···················· | ···················· | |
| V | 8687 hCoV-19/Portugal/PT1574/2020\|EPI_ISL_693582\|2020-0 | ···················· | ···················· | ···················· | ···················· | ···················· | ···················· | |
| V | 8688 hCoV-19/Portugal/PT1575/2020\|EPI_ISL_693583\|2020-0 | ···················· | ···················· | ···················· | ···················· | ···················· | ···················· | |
| V | 8689 hCoV-19/Portugal/PT1576/2020\|EPI_ISL_693584\|2020-0 | ···················· | ···················· | ···················· | ···················· | ··N····NN··········· | ···················· | 1 |
| V | 8690 hCoV-19/Portugal/PT1577/2020\|EPI_ISL_693585\|2020-0 | ···················· | ···················· | ···················· | ···················· | ···················· | ···················· | |
| V | 8691 hCoV-19/Portugal/PT1578/2020\|EPI_ISL_693586\|2020-0 | ···················· | ···················· | ···················· | ···················· | ···················· | ···················· | |
| V | 8692 hCoV-19/Portugal/PT1579/2020\|EPI_ISL_693587\|2020-0 | ···················· | ···················· | ···················· | ···················· | ···················· | ···················· | |
| V | 8693 hCoV-19/Portugal/PT1581/2020\|EPI_ISL_693588\|2020-0 | ···················· | ···················· | ···················· | ···················· | ···················· | ···················· | |
| V | 8694 hCoV-19/Portugal/PT1583/2020\|EPI_ISL_693589\|2020-0 | ···················· | ···················· | ···················· | ···················· | ···················· | ···················· | |
| V | 8698 hCoV-19/Portugal/PT1588/2020\|EPI_ISL_693593\|2020-0 | ···················· | ···················· | ···················· | ···················· | ···················· | ···················· | |
| V | 8700 hCoV-19/Portugal/PT1590/2020\|EPI_ISL_693595\|2020-0 | ···················· | ···················· | ···················· | ···················· | ···················· | ···················· | |
| V | 8702 hCoV-19/Portugal/PT1592/2020\|EPI_ISL_693597\|2020-0 | ···················· | ···················· | ···················· | ···················· | ···················· | ···················· | |
| V | 8703 hCoV-19/Portugal/PT1593/2020\|EPI_ISL_693598\|2020-0 | ···················· | ···················· | ···················· | ···················· | ···················· | ···················· | |
| V | 8705 hCoV-19/Japan/PG-1393/2020\|EPI_ISL_685000\|2020-04 | ···················· | ···················· | ···················· | ···················· | ···················· | ···················· | |
| V | 8706 hCoV-19/Japan/PG-1394/2020\|EPI_ISL_685001\|2020-04 | ···················· | ···················· | ···················· | ···················· | ···················· | ···················· | |
| V | 8707 hCoV-19/Japan/PG-1395/2020\|EPI_ISL_685002\|2020-04 | ···················· | ···················· | ···················· | ···················· | ···················· | ···················· | |
| V | 8708 hCoV-19/Japan/PG-1396/2020\|EPI_ISL_685003\|2020-04 | ···················· | ···················· | ···················· | ···················· | ···················· | ···················· | |
| V | 8709 hCoV-19/Japan/PG-1397/2020\|EPI_ISL_685004\|2020-04 | ···················· | ···················· | ···················· | ···················· | ···················· | ···················· | |
| V | 8710 hCoV-19/Japan/PG-1398/2020\|EPI_ISL_685005\|2020-04 | ···················· | ···················· | ···················· | ···················· | ···················· | ···················· | |
| V | 8711 hCoV-19/Japan/PG-1399/2020\|EPI_ISL_685006\|2020-04 | ···················· | ···················· | ···················· | ···················· | ···················· | ···················· | |
| V | 8712 hCoV-19/Japan/PG-1400/2020\|EPI_ISL_685007\|2020-04 | ···················· | ···················· | ···················· | ···················· | ···················· | ···················· | |
| V | 8713 hCoV-19/Japan/PG-1401/2020\|EPI_ISL_685008\|2020-04 | ···················· | ···················· | ···················· | ···················· | ···················· | ···················· | |
| V | 8714 hCoV-19/Japan/PG-1402/2020\|EPI_ISL_685009\|2020-04 | ···················· | ···················· | ···················· | ···················· | ···················· | ···················· | |
| V | 8715 hCoV-19/Japan/PG-1403/2020\|EPI_ISL_685010\|2020-04 | ···················· | ···················· | ···················· | ···················· | ···················· | ···················· | |
| V | 8716 hCoV-19/Japan/PG-1404/2020\|EPI_ISL_685011\|2020-04 | ···················· | ···················· | ···················· | ···················· | ···················· | ···················· | |
| V | 8717 hCoV-19/Japan/PG-1405/2020\|EPI_ISL_685012\|2020-04 | ···················· | ···················· | ···················· | ···················· | ···················· | ···················· | |
| V | 8718 hCoV-19/Japan/PG-1406/2020\|EPI_ISL_685013\|2020-04 | ···················· | ···················· | ···················· | ···················· | ···················· | ···················· | |
| V | 8719 hCoV-19/Japan/PG-1407/2020\|EPI_ISL_685014\|2020-04 | ···················· | ···················· | ···················· | ···················· | ···················· | ···················· | |
| V | 8720 hCoV-19/Japan/PG-1408/2020\|EPI_ISL_685015\|2020-04 | ···················· | ···················· | ···················· | ···················· | ···················· | ···················· | |
| V | 8721 hCoV-19/Japan/PG-1409/2020\|EPI_ISL_685016\|2020-04 | ···················· | ···················· | ···················· | ···················· | ···················· | ···················· | |
| V | 8722 hCoV-19/Japan/PG-1410/2020\|EPI_ISL_685017\|2020-04 | ···················· | ···················· | ···················· | ···················· | ···················· | ···················· | |
| V | 8723 hCoV-19/Japan/PG-1411/2020\|EPI_ISL_685018\|2020-04 | ···················· | ···················· | ···················· | ···················· | ···················· | ···················· | |
| V | 8724 hCoV-19/Japan/PG-1412/2020\|EPI_ISL_685019\|2020-04 | ···················· | ···················· | ···················· | ···················· | ···················· | ···················· | |
| V | 8725 hCoV-19/Japan/PG-1413/2020\|EPI_ISL_685020\|2020-04 | ···················· | ···················· | ···················· | ···················· | ···················· | ···················· | |
| V | 8726 hCoV-19/Japan/PG-1414/2020\|EPI_ISL_685021\|2020-04 | ···················· | ···················· | ···················· | ···················· | ·····C·············· | ···················· | |
| V | 8727 hCoV-19/Japan/PG-1415/2020\|EPI_ISL_685022\|2020-04 | ···················· | ···················· | ···················· | ···················· | ···················· | ···················· | |

| | | | | | | | |
|---|---|---|---|---|---|---|---|
| V | 8728 hCoV-19/Japan/PG-1416/2020\|EPI_ISL_685023\|2020-04 | ···················· | ···················· | ···················· | ···················· | ···················· | ···················· |
| V | 8729 hCoV-19/Japan/PG-1417/2020\|EPI_ISL_685024\|2020-04 | ···················· | ···················· | ···················· | ···················· | ···················· | ···················· |
| V | 8730 hCoV-19/Japan/PG-1418/2020\|EPI_ISL_685025\|2020-04 | ···················· | ···················· | ···················· | ···················· | ···················· | ···················· |
| V | 8731 hCoV-19/Japan/PG-1419/2020\|EPI_ISL_685026\|2020-04 | ···················· | ···················· | ···················· | ···················· | ···················· | ···················· |
| V | 8732 hCoV-19/Japan/PG-1420/2020\|EPI_ISL_685027\|2020-04 | ···················· | ···················· | ···················· | ···················· | ···················· | ···················· |
| V | 8733 hCoV-19/Japan/PG-1421/2020\|EPI_ISL_685028\|2020-04 | ···················· | ···················· | ···················· | ···················· | ···················· | ···················· |
| V | 8734 hCoV-19/Japan/PG-1422/2020\|EPI_ISL_685029\|2020-04 | ···················· | ···················· | ···················· | ···················· | ···················· | ···················· |
| V | 8735 hCoV-19/Japan/PG-1423/2020\|EPI_ISL_685030\|2020-04 | ···················· | ···················· | ···················· | ···················· | ···················· | ···················· |
| V | 8736 hCoV-19/Japan/PG-1424/2020\|EPI_ISL_685031\|2020-04 | ···················· | ···················· | ···················· | ···················· | ···················· | ···················· |
| V | 8737 hCoV-19/Japan/PG-1425/2020\|EPI_ISL_685032\|2020-04 | ···················· | ···················· | ···················· | ···················· | ···················· | ···················· |
| V | 8738 hCoV-19/Japan/PG-1426/2020\|EPI_ISL_685033\|2020-04 | ···················· | ···················· | ···················· | ···················· | ···················· | ···················· |
| V | 8739 hCoV-19/Japan/PG-1427/2020\|EPI_ISL_685034\|2020-04 | ···················· | ···················· | ···················· | ···················· | ···················· | ···················· |
| V | 8740 hCoV-19/Japan/PG-1428/2020\|EPI_ISL_685035\|2020-04 | ···················· | ···················· | ···················· | ···················· | ···················· | ···················· |
| V | 8741 hCoV-19/Japan/PG-1429/2020\|EPI_ISL_685036\|2020-04 | ···················· | ···················· | ···················· | ···················· | ···················· | ···················· |
| V | 8742 hCoV-19/Japan/PG-1430/2020\|EPI_ISL_685037\|2020-04 | ···················· | ···················· | ···················· | ···················· | ···················· | ···················· |
| V | 8743 hCoV-19/Japan/PG-1431/2020\|EPI_ISL_685038\|2020-04 | ···················· | ···················· | ···················· | ···················· | ···················· | ···················· |
| V | 8744 hCoV-19/Japan/PG-1432/2020\|EPI_ISL_685039\|2020-04 | ···················· | ···················· | ···················· | ···················· | ···················· | ···················· |
| V | 8745 hCoV-19/Japan/PG-1433/2020\|EPI_ISL_685040\|2020-04 | ···················· | ···················· | ···················· | ···················· | ···················· | ···················· |
| V | 8746 hCoV-19/Japan/PG-1434/2020\|EPI_ISL_685041\|2020-04 | ···················· | ···················· | ···················· | ···················· | ···················· | ···················· |
| V | 8747 hCoV-19/Japan/PG-1435/2020\|EPI_ISL_685042\|2020-04 | ···················· | ···················· | ···················· | ···················· | ···················· | ···················· |
| V | 8748 hCoV-19/Japan/PG-1436/2020\|EPI_ISL_685043\|2020-04 | ···················· | ···················· | ···················· | ···················· | ···················· | ···················· |
| V | 8749 hCoV-19/Japan/PG-1437/2020\|EPI_ISL_685044\|2020-04 | ···················· | ···················· | ···················· | ···················· | ···················· | ···················· |
| V | 8750 hCoV-19/Japan/PG-1438/2020\|EPI_ISL_685045\|2020-04 | ···················· | ···················· | ···················· | ···················· | ···················· | ···················· |
| V | 8751 hCoV-19/Japan/PG-1439/2020\|EPI_ISL_685046\|2020-04 | ···················· | ···················· | ···················· | ···················· | ···················· | ···················· |
| V | 8752 hCoV-19/Japan/PG-1440/2020\|EPI_ISL_685047\|2020-04 | ···················· | ···················· | ···················· | ···················· | ···················· | ···················· |
| V | 8753 hCoV-19/Japan/PG-1441/2020\|EPI_ISL_685048\|2020-04 | ···················· | ···················· | ···················· | ···················· | ···················· | ···················· |
| V | 8754 hCoV-19/Japan/PG-1442/2020\|EPI_ISL_685049\|2020-04 | ···················· | ···················· | ···················· | ···················· | ···················· | ···················· |
| V | 8755 hCoV-19/Japan/PG-1443/2020\|EPI_ISL_685050\|2020-04 | ···················· | ···················· | ···················· | ···················· | ···················· | ···················· |
| V | 8756 hCoV-19/Japan/PG-1444/2020\|EPI_ISL_685051\|2020-04 | ···················· | ···················· | ···················· | ···················· | ···················· | ···················· |
| V | 8757 hCoV-19/Japan/PG-1446/2020\|EPI_ISL_685053\|2020-04 | ···················· | ···················· | ···················· | ···················· | ···················· | ···················· |
| V | 8758 hCoV-19/Japan/PG-1483/2020\|EPI_ISL_685090\|2020-04 | ···················· | ···················· | ···················· | ···················· | ···················· | ···················· |
| V | 8759 hCoV-19/Japan/PG-1484/2020\|EPI_ISL_685091\|2020-04 | ···················· | ···················· | ···················· | ···················· | ···················· | ···················· |
| V | 8760 hCoV-19/Japan/PG-1485/2020\|EPI_ISL_685092\|2020-04 | ···················· | ···················· | ···················· | ···················· | ···················· | ···················· |
| V | 8761 hCoV-19/Japan/PG-1486/2020\|EPI_ISL_685093\|2020-04 | ···················· | ···················· | ···················· | ···················· | ···················· | ···················· |
| V | 8762 hCoV-19/Japan/PG-1487/2020\|EPI_ISL_685094\|2020-04 | ···················· | ···················· | ···················· | ···················· | ···················· | ···················· |
| V | 8763 hCoV-19/Japan/PG-1488/2020\|EPI_ISL_685095\|2020-04 | ···················· | ···················· | ···················· | ···················· | ···················· | ···················· |
| V | 8764 hCoV-19/Japan/PG-1489/2020\|EPI_ISL_685096\|2020-04 | ···················· | ···················· | ···················· | ···················· | ···················· | ···················· |
| V | 8765 hCoV-19/Japan/PG-1490/2020\|EPI_ISL_685097\|2020-04 | ···················· | ···················· | ···················· | ···················· | ···················· | ···················· |
| V | 8766 hCoV-19/Japan/PG-1491/2020\|EPI_ISL_685098\|2020-04 | ···················· | ···················· | ···················· | ···················· | ···················· | ···················· |
| V | 8767 hCoV-19/Japan/PG-1492/2020\|EPI_ISL_685099\|2020-04 | ···················· | ···················· | ···················· | ···················· | ···················· | ···················· |
| V | 8768 hCoV-19/Japan/PG-8727/2020\|EPI_ISL_686323\|2020-08 | ···················· | ···················· | ···················· | ···················· | ···················· | ···················· |
| V | 8769 hCoV-19/Japan/PG-8729/2020\|EPI_ISL_686325\|2020-08 | ···················· | ···················· | ···················· | ···················· | ···················· | ···················· |
| V | 8770 hCoV-19/Japan/PG-8730/2020\|EPI_ISL_686326\|2020-08 | ···················· | ···················· | ···················· | ···················· | ···················· | ···················· |
| V | 8771 hCoV-19/Japan/PG-8731/2020\|EPI_ISL_686327\|2020-08 | ···················· | ···················· | ···················· | ···················· | ···················· | ···················· |
| V | 8772 hCoV-19/Japan/PG-8732/2020\|EPI_ISL_686328\|2020-08 | ···················· | ···················· | ···················· | ···················· | ···················· | ···················· |
| V | 8773 hCoV-19/Japan/PG-8733/2020\|EPI_ISL_686329\|2020-08 | ···················· | ···················· | ···················· | ···················· | ···················· | ···················· |
| V | 8774 hCoV-19/Japan/PG-8734/2020\|EPI_ISL_686330\|2020-08 | ···················· | ···················· | ···················· | ···················· | ···················· | ···················· |
| V | 8775 hCoV-19/Japan/PG-8735/2020\|EPI_ISL_686331\|2020-08 | ···················· | ···················· | ···················· | ···················· | ···················· | ···················· |
| V | 8776 hCoV-19/Japan/PG-8736/2020\|EPI_ISL_686332\|2020-08 | ···················· | ···················· | ···················· | ···················· | ···················· | ···················· |
| V | 8777 hCoV-19/Japan/PG-8737/2020\|EPI_ISL_686333\|2020-08 | ···················· | ···················· | ···················· | ···················· | ···················· | ···················· |
| V | 8778 hCoV-19/Japan/PG-8738/2020\|EPI_ISL_686334\|2020-08 | ···················· | ···················· | ···················· | ···················· | ···················· | ···················· |
| V | 8779 hCoV-19/Japan/PG-8739/2020\|EPI_ISL_686335\|2020-08 | ···················· | ···················· | ···················· | ···················· | ···················· | ···················· |
| V | 8780 hCoV-19/Japan/PG-8740/2020\|EPI_ISL_686336\|2020-09 | ···················· | ···················· | ···················· | ···················· | ···················· | ···················· |
| V | 8781 hCoV-19/Japan/PG-8741/2020\|EPI_ISL_686337\|2020-09 | ···················· | ···················· | ···················· | ···················· | ···················· | ···················· |
| V | 8782 hCoV-19/Japan/PG-8742/2020\|EPI_ISL_686338\|2020-09 | ···················· | ···················· | ···················· | ···················· | ···················· | ···················· |
| V | 8783 hCoV-19/Japan/PG-8743/2020\|EPI_ISL_686339\|2020-09 | ···················· | ···················· | ···················· | ···················· | ···················· | ···················· |
| V | 8784 hCoV-19/Japan/PG-8744/2020\|EPI_ISL_686340\|2020-09 | ···················· | ···················· | ···················· | ···················· | ···················· | ···················· |
| V | 8785 hCoV-19/Japan/PG-8745/2020\|EPI_ISL_686341\|2020-09 | ········T··········· | ···················· | ···················· | ···················· | ···················· | ···················· |
| V | 8786 hCoV-19/Japan/PG-8746/2020\|EPI_ISL_686342\|2020-09 | ···················· | ···················· | ···················· | ···················· | ···················· | ···················· |
| V | 8787 hCoV-19/Japan/PG-8747/2020\|EPI_ISL_686343\|2020-09 | ········T··········· | ···················· | ···················· | ···················· | ···················· | ···················· |
| V | 8788 hCoV-19/Japan/PG-8748/2020\|EPI_ISL_686344\|2020-09 | ···················· | ···················· | ···················· | ···················· | ···················· | ···················· |
| V | 8789 hCoV-19/Japan/PG-8749/2020\|EPI_ISL_686345\|2020-09 | ···················· | ······A············· | ···················· | ···················· | ···················· | ···················· |
| V | 8790 hCoV-19/Japan/PG-8750/2020\|EPI_ISL_686346\|2020-09 | ···················· | ···················· | ···················· | ···················· | ···················· | ···················· |
| V | 8791 hCoV-19/Japan/PG-8751/2020\|EPI_ISL_686347\|2020-09 | ···················· | ···················· | ···················· | ···················· | ···················· | ···················· |
| V | 8792 hCoV-19/Japan/PG-8752/2020\|EPI_ISL_686348\|2020-09 | ···················· | ···················· | ···················· | ···················· | ···················· | ···················· |
| V | 8793 hCoV-19/Japan/PG-8753/2020\|EPI_ISL_686349\|2020-09 | ···················· | ···················· | ···················· | ···················· | ···················· | ···················· |
| V | 8794 hCoV-19/Japan/PG-8754/2020\|EPI_ISL_686350\|2020-09 | ···················· | ···················· | ···················· | ···················· | ···················· | ···················· |
| V | 8795 hCoV-19/Japan/PG-8755/2020\|EPI_ISL_686351\|2020-09 | ···················· | ···················· | ···················· | ···················· | ···················· | ···················· |
| V | 8796 hCoV-19/Japan/PG-8756/2020\|EPI_ISL_686352\|2020-09 | ···················· | ···················· | ···················· | ···················· | ···················· | ···················· |
| V | 8797 hCoV-19/Japan/PG-8757/2020\|EPI_ISL_686353\|2020-09 | ···················· | ···················· | ···················· | ···················· | ···················· | ···················· |
| V | 8798 hCoV-19/Japan/PG-8758/2020\|EPI_ISL_686354\|2020-09 | ···················· | ···················· | ···················· | ···················· | ···················· | ···················· |
| V | 8799 hCoV-19/Japan/PG-8759/2020\|EPI_ISL_686355\|2020-09 | ···················· | ···················· | ···················· | ···················· | ···················· | ···················· |
| V | 8800 hCoV-19/Japan/PG-8760/2020\|EPI_ISL_686356\|2020-09 | ···················· | ···················· | ···················· | ···················· | ···················· | ···················· |
| V | 8801 hCoV-19/Japan/PG-8761/2020\|EPI_ISL_686357\|2020-09 | ········T··········· | ···················· | ···················· | ···················· | ···················· | ···················· |
| V | 8802 hCoV-19/Japan/PG-8762/2020\|EPI_ISL_686358\|2020-09 | ···················· | ···················· | ···················· | ···················· | ···················· | ···················· |
| V | 8803 hCoV-19/Japan/PG-8763/2020\|EPI_ISL_686359\|2020-09 | ···················· | ···················· | ···················· | ···················· | ···················· | ···················· |
| V | 8804 hCoV-19/Japan/PG-8764/2020\|EPI_ISL_686360\|2020-09 | ···················· | ···················· | ···················· | ···················· | ···················· | ···················· |
| V | 8805 hCoV-19/Japan/PG-8765/2020\|EPI_ISL_686361\|2020-09 | ········T··········· | ···················· | ···················· | ···················· | ···················· | ···················· |
| V | 8806 hCoV-19/Japan/PG-8766/2020\|EPI_ISL_686362\|2020-09 | ···················· | ···················· | ···················· | ···················· | ···················· | ···················· |
| V | 8807 hCoV-19/Japan/PG-8767/2020\|EPI_ISL_686363\|2020-09 | ···················· | ··········A········· | ···················· | ···················· | ···················· | ···················· |

| | | | | | | | | |
|---|---|---|---|---|---|---|---|---|
| V | 8808 | hCoV-19/Japan/PG-8768/2020\|EPI_ISL_686364\|2020-09 | .................... | .................... | ..................... | .................... | .................... | .................... |
| V | 8809 | hCoV-19/Japan/PG-8769/2020\|EPI_ISL_686365\|2020-09 | .................... | .................... | ..................... | .................... | .................... | .................... |
| V | 8810 | hCoV-19/Japan/PG-8770/2020\|EPI_ISL_686366\|2020-09 | .................... | .................... | ..................... | .................... | .................... | .................... |
| V | 8811 | hCoV-19/Japan/PG-8773/2020\|EPI_ISL_686367\|2020-08 | .................... | .................... | ..................... | .................... | .................... | .................... |
| V | 8812 | hCoV-19/Japan/PG-8774/2020\|EPI_ISL_686368\|2020-08 | .................... | .................... | ..................... | .................... | .................... | .................... |
| V | 8813 | hCoV-19/Japan/PG-8775/2020\|EPI_ISL_686369\|2020-08 | .................... | .................... | ..................... | .................... | .................... | .................... |
| V | 8814 | hCoV-19/Japan/PG-8776/2020\|EPI_ISL_686370\|2020-08 | .................... | .................... | ..................... | .................... | .................... | .................... |
| V | 8815 | hCoV-19/Japan/PG-8777/2020\|EPI_ISL_686371\|2020-08 | .................... | .................... | ..................... | .................... | .................... | .................... |
| V | 8816 | hCoV-19/Japan/PG-8781/2020\|EPI_ISL_686372\|2020-08 | .................... | .................... | ..................... | .................... | .................... | .................... |
| V | 8817 | hCoV-19/Japan/PG-8782/2020\|EPI_ISL_686373\|2020-08 | .................... | .................... | ..................... | .................... | .................... | .................... |
| V | 8818 | hCoV-19/Japan/PG-8783/2020\|EPI_ISL_686374\|2020-08 | .................... | .................... | ..................... | .................... | .................... | .................... |
| V | 8819 | hCoV-19/Japan/PG-8785/2020\|EPI_ISL_686375\|2020-08 | .................... | .................... | ..................... | .................... | .................... | .................... |
| V | 8820 | hCoV-19/Japan/PG-8787/2020\|EPI_ISL_686376\|2020-08 | .................... | .................... | ..................... | .................... | .................... | .................... |
| V | 8821 | hCoV-19/Japan/PG-8788/2020\|EPI_ISL_686377\|2020-08 | .................... | .................... | ..................... | .................... | .................... | .................... |
| V | 8822 | hCoV-19/Japan/PG-8789/2020\|EPI_ISL_686378\|2020-08 | .................... | .................... | ..................... | .................... | .................... | .................... |
| V | 8823 | hCoV-19/Japan/PG-8791/2020\|EPI_ISL_686379\|2020-08 | .................... | .................... | ..................... | .................... | .................... | .................... |
| V | 8824 | hCoV-19/Japan/PG-8792/2020\|EPI_ISL_686380\|2020-08 | .................... | .................... | ..................... | .................... | .................... | .................... |
| V | 8825 | hCoV-19/Japan/PG-8793/2020\|EPI_ISL_686381\|2020-08 | .................... | .................... | ..................... | .................... | .................... | .................... |
| V | 8826 | hCoV-19/Japan/PG-8794/2020\|EPI_ISL_686382\|2020-08 | .................... | .................... | ..................... | .................... | .................... | .................... |
| V | 8827 | hCoV-19/Japan/PG-8796/2020\|EPI_ISL_686384\|2020-08 | .................... | .................... | ..................... | .................... | .................... | .................... |
| V | 8828 | hCoV-19/Japan/PG-6541/2020\|EPI_ISL_687655\|2020-07 | .................... | .................... | ..................... | .................... | .................... | .................... |
| V | 8829 | hCoV-19/Japan/PG-6542/2020\|EPI_ISL_687656\|2020-08 | .................... | .................... | ..................... | .................... | .................... | .................... |
| V | 8830 | hCoV-19/Japan/PG-6543/2020\|EPI_ISL_687657\|2020-08 | .................... | .................... | ..................... | .................... | .................... | .................... |
| V | 8831 | hCoV-19/Japan/PG-6544/2020\|EPI_ISL_687658\|2020-08 | .................... | .................... | ..................... | .................... | .................... | .................... |
| V | 8832 | hCoV-19/Japan/PG-6545/2020\|EPI_ISL_687659\|2020-08 | .................... | .................... | ..................... | .................... | .................... | .................... |
| V | 8833 | hCoV-19/Japan/PG-6546/2020\|EPI_ISL_687660\|2020-08 | .................... | .................... | ..................... | .................... | .................... | .................... |
| V | 8834 | hCoV-19/Japan/PG-6547/2020\|EPI_ISL_687661\|2020-08 | .................... | .................... | ..................... | .................... | .................... | .................... |
| V | 8835 | hCoV-19/Japan/PG-6548/2020\|EPI_ISL_687662\|2020-08 | .................... | .................... | ..................... | .................... | .................... | .................... |
| V | 8836 | hCoV-19/Japan/PG-6549/2020\|EPI_ISL_687663\|2020-08 | .................... | .................... | ..................... | .................... | .................... | .................... |
| V | 8837 | hCoV-19/Japan/PG-6550/2020\|EPI_ISL_687664\|2020-08 | .................... | .................... | ..................... | .................... | .................... | .................... |
| V | 8838 | hCoV-19/Japan/PG-6551/2020\|EPI_ISL_687665\|2020-08 | .................... | .................... | ..................... | .................... | .................... | .................... |
| V | 8839 | hCoV-19/Japan/PG-6552/2020\|EPI_ISL_687666\|2020-08 | .................... | .................... | ..................... | .................... | .................... | .................... |
| V | 8840 | hCoV-19/Japan/PG-6553/2020\|EPI_ISL_687667\|2020-08 | .................... | .................... | ..................... | .................... | .................... | .................... |
| V | 8841 | hCoV-19/Japan/PG-6554/2020\|EPI_ISL_687668\|2020-08 | .................... | .................... | ..................... | .................... | .................... | .................... |
| V | 8842 | hCoV-19/Japan/PG-6555/2020\|EPI_ISL_687669\|2020-09 | .................... | .................... | ..................... | .................... | .................... | .................... |
| V | 8843 | hCoV-19/Japan/PG-6556/2020\|EPI_ISL_687670\|2020-09 | .................... | .................... | ..................... | .................... | .................... | .................... |
| V | 8844 | hCoV-19/Japan/PG-6557/2020\|EPI_ISL_687671\|2020-09 | .................... | .................... | ..................... | .................... | .................... | .................... |
| V | 8845 | hCoV-19/Japan/PG-6558/2020\|EPI_ISL_687672\|2020-09 | .................... | .................... | ..................... | .................... | .................... | .................... |
| V | 8846 | hCoV-19/Japan/PG-6559/2020\|EPI_ISL_687673\|2020-08 | .................... | .................... | ..................... | .................... | .................... | .................... |
| V | 8847 | hCoV-19/Japan/PG-6560/2020\|EPI_ISL_687674\|2020-08 | .................... | .................... | ..................... | .................... | .................... | .................... |
| V | 8848 | hCoV-19/Japan/PG-6561/2020\|EPI_ISL_687675\|2020-08 | .................... | .................... | ..................... | .................... | .................... | .................... |
| V | 8849 | hCoV-19/Japan/PG-6562/2020\|EPI_ISL_687676\|2020-09 | .................... | .................... | ..................... | .................... | .................... | .................... |
| V | 8850 | hCoV-19/Japan/PG-6565/2020\|EPI_ISL_687677\|2020-09 | .................... | .................... | ..................... | .................... | .................... | .................... |
| V | 8851 | hCoV-19/Japan/PG-6566/2020\|EPI_ISL_687678\|2020-09 | .................... | .................... | ..................... | .................... | .................... | .................... |
| V | 8852 | hCoV-19/Japan/PG-6567/2020\|EPI_ISL_687679\|2020-09 | .................... | .................... | ..................... | .................... | .................... | .................... |
| V | 8853 | hCoV-19/Japan/PG-6568/2020\|EPI_ISL_687680\|2020-09 | .................... | .................... | ..................... | .................... | .................... | .................... |
| V | 8854 | hCoV-19/Japan/PG-6569/2020\|EPI_ISL_687681\|2020-09 | .................... | .................... | ..................... | .................... | .................... | .................... |
| V | 8855 | hCoV-19/Japan/PG-6570/2020\|EPI_ISL_687682\|2020-09 | .................... | .................... | ..................... | .................... | .................... | .................... |
| V | 8856 | hCoV-19/Japan/PG-6571/2020\|EPI_ISL_687683\|2020-09 | .................... | .................... | ..................... | .................... | .................... | .................... |
| V | 8857 | hCoV-19/Japan/PG-6572/2020\|EPI_ISL_687684\|2020-09 | .................... | .................... | ..................... | .................... | .................... | .................... |
| V | 8858 | hCoV-19/Japan/PG-6573/2020\|EPI_ISL_687685\|2020-09 | .................... | .................... | ..................... | .................... | .................... | .................... |
| V | 8859 | hCoV-19/Japan/PG-6574/2020\|EPI_ISL_687686\|2020-09 | .................... | .................... | ..................... | .................... | .................... | .................... |
| V | 8860 | hCoV-19/Japan/PG-6575/2020\|EPI_ISL_687687\|2020-09 | .................... | .................... | ..................... | .................... | .................... | .................... |
| V | 8861 | hCoV-19/Japan/PG-6576/2020\|EPI_ISL_687688\|2020-09 | .................... | .................... | ..................... | .................... | .................... | .................... |
| V | 8862 | hCoV-19/Japan/PG-6577/2020\|EPI_ISL_687689\|2020-09 | .................... | .................... | ..................... | .................... | .................... | .................... |
| V | 8863 | hCoV-19/Japan/PG-6578/2020\|EPI_ISL_687690\|2020-09 | .................... | .................... | ..................... | .................... | .................... | .................... |
| V | 8864 | hCoV-19/Japan/PG-6579/2020\|EPI_ISL_687691\|2020-09 | .................... | .................... | ..................... | .................... | .................... | .................... |
| V | 8865 | hCoV-19/Japan/PG-6580/2020\|EPI_ISL_687692\|2020-09 | .................... | .................... | ..................... | .................... | .................... | .................... |
| V | 8866 | hCoV-19/Japan/PG-6581/2020\|EPI_ISL_687693\|2020-09 | .................... | .................... | ..................... | .................... | .................... | .................... |
| V | 8867 | hCoV-19/Japan/PG-6582/2020\|EPI_ISL_687694\|2020-09 | .................... | .................... | ..................... | .................... | .................... | .................... |
| V | 8868 | hCoV-19/Japan/PG-6583/2020\|EPI_ISL_687695\|2020-09 | .................... | .................... | ..................... | .................... | .................... | .................... |
| V | 8869 | hCoV-19/Japan/PG-6584/2020\|EPI_ISL_687696\|2020-09 | .................... | .................... | ..................... | .................... | .................... | .................... |
| V | 8870 | hCoV-19/Japan/PG-6585/2020\|EPI_ISL_687697\|2020-09 | .................... | .................... | ..................... | .................... | .................... | .................... |
| V | 8871 | hCoV-19/Japan/PG-6586/2020\|EPI_ISL_687698\|2020-09 | .................... | .................... | ..................... | .................... | .................... | .................... |
| V | 8872 | hCoV-19/Japan/PG-6587/2020\|EPI_ISL_687699\|2020-07 | .................... | .................... | ..................... | .................... | .................... | .................... |
| V | 8873 | hCoV-19/Japan/PG-5781/2020\|EPI_ISL_688900\|2020-08 | .................... | .................... | ..................... | .................... | .................... | .................... |
| V | 8874 | hCoV-19/Japan/PG-5782/2020\|EPI_ISL_688901\|2020-08 | .................... | .................... | ..................... | .................... | .................... | .................... |
| V | 8875 | hCoV-19/Japan/PG-5783/2020\|EPI_ISL_688902\|2020-08 | .................... | .................... | ..................... | .................... | .................... | .................... |
| V | 8876 | hCoV-19/Japan/PG-5784/2020\|EPI_ISL_688903\|2020-08 | .................... | .................... | ..................... | .................... | .................... | .................... |
| V | 8877 | hCoV-19/Japan/PG-5785/2020\|EPI_ISL_688904\|2020-08 | .................... | .................... | ..................... | .................... | .................... | .................... |
| V | 8878 | hCoV-19/Japan/PG-5786/2020\|EPI_ISL_688905\|2020-08 | .................... | .................... | ..................... | .................... | .................... | .................... |
| V | 8879 | hCoV-19/Japan/PG-5787/2020\|EPI_ISL_688906\|2020-08 | .................... | .................... | ..................... | .................... | .................... | .................... |
| V | 8880 | hCoV-19/Japan/PG-5788/2020\|EPI_ISL_688907\|2020-08 | .................... | .................... | ..................... | .................... | .................... | .................... |
| V | 8881 | hCoV-19/Japan/PG-5789/2020\|EPI_ISL_688908\|2020-08 | .................... | .................... | ..................... | .................... | .................... | .................... |
| V | 8882 | hCoV-19/Japan/PG-5790/2020\|EPI_ISL_688909\|2020-08 | .................... | .................... | ..................... | .................... | .................... | .................... |
| V | 8883 | hCoV-19/Japan/PG-5871/2020\|EPI_ISL_688986\|2020-08 | .................... | .................... | ..................... | .................... | .................... | .................... |
| V | 8884 | hCoV-19/Japan/PG-5872/2020\|EPI_ISL_688987\|2020-08 | .................... | .................... | ..................... | .................... | .................... | .................... |
| V | 8885 | hCoV-19/Japan/PG-5873/2020\|EPI_ISL_688988\|2020-08 | .................... | .................... | ..................... | .................... | .................... | .................... |
| V | 8886 | hCoV-19/Japan/PG-5874/2020\|EPI_ISL_688989\|2020-08 | .................... | .................... | ..................... | .................... | .................... | .................... |
| V | 8887 | hCoV-19/Japan/PG-5875/2020\|EPI_ISL_688990\|2020-08 | .................... | .................... | ..................... | .................... | .................... | .................... |

| | | | | | | | | |
|---|---|---|---|---|---|---|---|---|
| V | 8888 | hCoV-19/Japan/PG-5876/2020\|EPI_ISL_688991\|2020-08 | ···················· | ···················· | ····················· | ···················· | ···················· | ···················· |
| V | 8889 | hCoV-19/Japan/PG-5877/2020\|EPI_ISL_688992\|2020-08 | ···················· | ···················· | ····················· | ···················· | ···················· | ···················· |
| V | 8890 | hCoV-19/Japan/PG-5878/2020\|EPI_ISL_688993\|2020-08 | ···················· | ···················· | ····················· | ···················· | ···················· | ···················· |
| V | 8891 | hCoV-19/Japan/PG-5879/2020\|EPI_ISL_688994\|2020-08 | ···················· | ···················· | ····················· | ···················· | ···················· | ···················· |
| V | 8892 | hCoV-19/Japan/PG-5880/2020\|EPI_ISL_688995\|2020-08 | ···················· | ···················· | ····················· | ···················· | ···················· | ···················· |
| V | 8893 | hCoV-19/Japan/PG-5881/2020\|EPI_ISL_688996\|2020-08 | ···················· | ···················· | ····················· | ···················· | ···················· | ···················· |
| V | 8894 | hCoV-19/Japan/PG-5882/2020\|EPI_ISL_688997\|2020-08 | ···················· | ···················· | ····················· | ···················· | ···················· | ···················· |
| V | 8895 | hCoV-19/Japan/PG-5883/2020\|EPI_ISL_688998\|2020-08 | ···················· | ···················· | ····················· | ···················· | ···················· | ···················· |
| V | 8896 | hCoV-19/Japan/PG-5884/2020\|EPI_ISL_688999\|2020-08 | ···················· | ···················· | ····················· | ···················· | ···················· | ···················· |
| V | 8897 | hCoV-19/Japan/PG-3926/2020\|EPI_ISL_692177\|2020-07 | ···················· | ···················· | ····················· | ···················· | ···················· | ···················· |
| V | 8898 | hCoV-19/Japan/PG-3927/2020\|EPI_ISL_692178\|2020-07 | ···················· | ···················· | ····················· | ···················· | ···················· | ···················· |
| V | 8899 | hCoV-19/Japan/PG-3928/2020\|EPI_ISL_692179\|2020-07 | ···················· | ···················· | ····················· | ···················· | ···················· | ···················· |
| V | 8900 | hCoV-19/Japan/PG-3929/2020\|EPI_ISL_692180\|2020-07 | ···················· | ···················· | ····················· | ···················· | ···················· | ···················· |
| V | 8901 | hCoV-19/Japan/PG-3930/2020\|EPI_ISL_692181\|2020-07 | ···················· | ···················· | ····················· | ···················· | ···················· | ···················· |
| V | 8902 | hCoV-19/Japan/PG-3931/2020\|EPI_ISL_692182\|2020-07 | ···················· | ···················· | ····················· | ···················· | ···················· | ···················· |
| V | 8903 | hCoV-19/Japan/PG-3932/2020\|EPI_ISL_692183\|2020-07 | ···················· | ···················· | ····················· | ···················· | ···················· | ···················· |
| V | 8904 | hCoV-19/Japan/PG-3933/2020\|EPI_ISL_692184\|2020-07 | ···················· | ···················· | ····················· | ···················· | ···················· | ···················· |
| V | 8905 | hCoV-19/Japan/PG-3934/2020\|EPI_ISL_692185\|2020-07 | ···················· | ···················· | ····················· | ···················· | ···················· | ···················· |
| V | 8906 | hCoV-19/Japan/PG-3935/2020\|EPI_ISL_692186\|2020-07 | ···················· | ···················· | ····················· | ···················· | ···················· | ···················· |
| V | 8907 | hCoV-19/Japan/PG-3936/2020\|EPI_ISL_692187\|2020-07 | ···················· | ···················· | ····················· | ···················· | ···················· | ···················· |
| V | 8908 | hCoV-19/Japan/PG-3937/2020\|EPI_ISL_692188\|2020-07 | ···················· | ···················· | ····················· | ···················· | ···················· | ···················· |
| V | 8909 | hCoV-19/Japan/PG-3938/2020\|EPI_ISL_692189\|2020-07 | ···················· | ···················· | ····················· | ···················· | ···················· | ···················· |
| V | 8910 | hCoV-19/Japan/PG-3941/2020\|EPI_ISL_692190\|2020-07 | ···················· | ···················· | ····················· | ···················· | ···················· | ···················· |
| V | 8911 | hCoV-19/Japan/PG-3942/2020\|EPI_ISL_692191\|2020-06 | ···················· | ···················· | ····················· | ···················· | ···················· | ···················· |
| V | 8912 | hCoV-19/Japan/PG-3943/2020\|EPI_ISL_692192\|2020-07 | ···················· | ···················· | ····················· | ···················· | ···················· | ···················· |
| V | 8913 | hCoV-19/Japan/PG-3944/2020\|EPI_ISL_692193\|2020-07 | ···················· | ···················· | ····················· | ···················· | ···················· | ···················· |
| V | 8914 | hCoV-19/Japan/PG-3945/2020\|EPI_ISL_692194\|2020-07 | ···················· | ···················· | ····················· | ···················· | ···················· | ···················· |
| V | 8915 | hCoV-19/Japan/PG-3946/2020\|EPI_ISL_692195\|2020-07 | ···················· | ···················· | ····················· | ···················· | ···················· | ···················· |
| V | 8917 | hCoV-19/Japan/PG-3948/2020\|EPI_ISL_692197\|2020-04 | ···················· | ···················· | ····················· | ···················· | ···················· | ···················· |
| V | 8918 | hCoV-19/Japan/PG-3949/2020\|EPI_ISL_692198\|2020-07 | ···················· | ···················· | ····················· | ···················· | ···················· | ···················· |
| V | 8919 | hCoV-19/Japan/PG-3950/2020\|EPI_ISL_692199\|2020-07 | ···················· | ···················· | ····················· | ···················· | ···················· | ···················· |
| V | 8920 | hCoV-19/Japan/PG-3951/2020\|EPI_ISL_692200\|2020-07 | ···················· | ···················· | ····················· | ···················· | ···················· | ···················· |
| V | 8921 | hCoV-19/Japan/PG-3952/2020\|EPI_ISL_692201\|2020-07 | ···················· | ···················· | ····················· | ···················· | ···················· | ···················· |
| V | 8922 | hCoV-19/Japan/PG-3953/2020\|EPI_ISL_692202\|2020-07 | ···················· | ···················· | ····················· | ···················· | ···················· | ···················· |
| V | 8923 | hCoV-19/Japan/PG-3954/2020\|EPI_ISL_692203\|2020-07 | ···················· | ···················· | ····················· | ···················· | ···················· | ···················· |
| V | 8924 | hCoV-19/Japan/PG-3955/2020\|EPI_ISL_692204\|2020-07 | ···················· | ···················· | ····················· | ···················· | ···················· | ···················· |
| V | 8925 | hCoV-19/Japan/PG-3956/2020\|EPI_ISL_692205\|2020-07 | ···················· | ···················· | ····················· | ···················· | ···················· | ···················· |
| V | 8926 | hCoV-19/Japan/PG-3958/2020\|EPI_ISL_692206\|2020-07 | ···················· | ···················· | ····················· | ···················· | ···················· | ···················· |
| V | 8927 | hCoV-19/Japan/PG-3959/2020\|EPI_ISL_692207\|2020-07 | ···················· | ···················· | ····················· | ···················· | ···················· | ···················· |
| V | 8928 | hCoV-19/Japan/PG-3960/2020\|EPI_ISL_692208\|2020-07 | ···················· | ···················· | ····················· | ···················· | ···················· | ···················· |
| V | 8929 | hCoV-19/Japan/PG-3961/2020\|EPI_ISL_692209\|2020-07 | ···················· | ···················· | ····················· | ···················· | ···················· | ···················· |
| V | 8930 | hCoV-19/England/204682076/2020\|EPI_ISL_693420\|2020 | ···················· | ···················· | ····················· | ···················· | ···················· | ···················· |
| V | 8931 | hCoV-19/England/204682079/2020\|EPI_ISL_693421\|2020 | ···················· | ·A·················· | ····················· | ···················· | ···················· | ···················· |
| V | 8932 | hCoV-19/England/204682081/2020\|EPI_ISL_693422\|2020 | ···················· | ···················· | ····················· | ···················· | ···················· | ···················· |
| V | 8933 | hCoV-19/England/204682089/2020\|EPI_ISL_693423\|2020 | ···················· | ···················· | ····················· | ···················· | ···················· | ···················· |
| V | 8934 | hCoV-19/England/204682404/2020\|EPI_ISL_693424\|2020 | ···················· | ···················· | ····················· | ···················· | ···················· | ···················· |
| V | 8935 | hCoV-19/England/204682408/2020\|EPI_ISL_693425\|2020 | ···················· | ···················· | ····················· | ···················· | ···················· | ···················· |
| V | 8936 | hCoV-19/England/204682412/2020\|EPI_ISL_693426\|2020 | ···················· | ···················· | ····················· | ···················· | ···················· | ···················· |
| V | 8937 | hCoV-19/England/204682424/2020\|EPI_ISL_693427\|2020 | ···················· | ···················· | ····················· | ···················· | ···················· | ···················· |
| V | 8938 | hCoV-19/England/204683182/2020\|EPI_ISL_693428\|2020 | ···················· | ···················· | ····················· | ···················· | ···················· | ···················· |
| V | 8939 | hCoV-19/England/204683193/2020\|EPI_ISL_693429\|2020 | ···················· | ···················· | ····················· | ···················· | ·····T·············· | ···················· |
| V | 8940 | hCoV-19/England/204683196/2020\|EPI_ISL_693430\|2020 | ···················· | ···················· | ····················· | ···················· | ···················· | ···················· |
| V | 8941 | hCoV-19/France/ARA-74594/2020\|EPI_ISL_693500\|2020- | ···················· | ···················· | ····················· | ···················· | ···················· | ···················· |
| V | 8942 | hCoV-19/France/ARA-75471/2020\|EPI_ISL_693501\|2020- | ···················· | ···················· | ····················· | ···················· | ···················· | ···················· |
| V | 8943 | hCoV-19/France/ARA-75858/2020\|EPI_ISL_693502\|2020- | ···················· | ···················· | ····················· | ···················· | ···················· | ···················· |
| V | 8944 | hCoV-19/France/ARA-76483/2020\|EPI_ISL_693503\|2020- | ···················· | ···················· | ····················· | ···················· | ···················· | ···················· |
| V | 8945 | hCoV-19/France/ARA-76531/2020\|EPI_ISL_693504\|2020- | ···················· | ···················· | ····················· | ···················· | ···················· | ···················· |
| V | 8946 | hCoV-19/France/ARA-76532/2020\|EPI_ISL_693505\|2020- | ···················· | ···················· | ····················· | ···················· | ···················· | ···················· |
| V | 8947 | hCoV-19/France/ARA-76738/2020\|EPI_ISL_693506\|2020- | ···················· | ···················· | ····················· | ···················· | ···················· | ···················· |
| V | 8948 | hCoV-19/France/ARA-77036/2020\|EPI_ISL_693507\|2020- | ···················· | ···················· | ····················· | ···················· | ···················· | ···················· |
| V | 8949 | hCoV-19/France/ARA-77064/2020\|EPI_ISL_693508\|2020- | ···················· | ···················· | ···A················· | ···················· | ···················· | ···················· |
| V | 8950 | hCoV-19/France/ARA-77159/2020\|EPI_ISL_693509\|2020- | ···················· | ···················· | ····················· | ···················· | ···················· | ···················· |
| V | 8951 | hCoV-19/France/ARA-77597/2020\|EPI_ISL_693510\|2020- | ···················· | ···················· | ····················· | ···················· | ···················· | ···················· |
| V | 8952 | hCoV-19/France/ARA-77679/2020\|EPI_ISL_693511\|2020- | ···················· | ···················· | ····················· | ···················· | ···················· | ···················· |
| V | 8953 | hCoV-19/France/ARA-78039/2020\|EPI_ISL_693512\|2020- | ···················· | ···················· | ····················· | ···················· | ···················· | ···················· |
| V | 8954 | hCoV-19/France/ARA-78043/2020\|EPI_ISL_693513\|2020- | ···················· | ···················· | ····················· | ···················· | ···················· | ···················· |
| V | 8955 | hCoV-19/France/ARA-78085/2020\|EPI_ISL_693514\|2020- | ···················· | ···················· | ····················· | ···················· | ···················· | ···················· |
| V | 8956 | hCoV-19/Portugal/PT1557/2020\|EPI_ISL_693529\|2020-0 | ···················· | ···················· | ····················· | ···················· | ···················· | ···················· |
| V | 8957 | hCoV-19/Portugal/PT1582/2020\|EPI_ISL_693530\|2020-0 | ···················· | ···················· | ····················· | ···················· | ···················· | ···················· |
| V | 8958 | hCoV-19/Portugal/PT1609/2020\|EPI_ISL_693531\|2020-0 | ···················· | ···················· | ····················· | ···················· | ···················· | ···················· |
| V | 8959 | hCoV-19/Portugal/PT1626/2020\|EPI_ISL_693532\|2020-0 | ···················· | ···················· | ····················· | ···················· | ···················· | ···················· |
| V | 8960 | hCoV-19/Portugal/PT1580/2020\|EPI_ISL_693533\|2020-0 | ···················· | ···················· | ····················· | ···················· | ···················· | ···················· |
| V | 8961 | hCoV-19/Japan/PG-8704/2020\|EPI_ISL_686300\|2020-08 | ···················· | ···················· | ····················· | ···················· | ···················· | ···················· |
| V | 8962 | hCoV-19/Japan/PG-8705/2020\|EPI_ISL_686301\|2020-08 | ···················· | ···················· | ····················· | ···················· | ···················· | ···················· |
| V | 8963 | hCoV-19/Japan/PG-8706/2020\|EPI_ISL_686302\|2020-08 | ···················· | ···················· | ····················· | ···················· | ···················· | ···················· |
| V | 8964 | hCoV-19/Japan/PG-8707/2020\|EPI_ISL_686303\|2020-08 | ···················· | ···················· | ····················· | ···················· | ···················· | ···················· |
| V | 8965 | hCoV-19/Japan/PG-8708/2020\|EPI_ISL_686304\|2020-08 | ···················· | ···················· | ····················· | ···················· | ···················· | ···················· |
| V | 8966 | hCoV-19/Japan/PG-8709/2020\|EPI_ISL_686305\|2020-08 | ···················· | ···················· | ····················· | ···················· | ···················· | ···················· |
| V | 8967 | hCoV-19/Japan/PG-8710/2020\|EPI_ISL_686306\|2020-08 | ···················· | ···················· | ····················· | ···················· | ···················· | ···················· |
| V | 8968 | hCoV-19/Japan/PG-8711/2020\|EPI_ISL_686307\|2020-08 | ···················· | ···················· | ····················· | ···················· | ···················· | ···················· |

| | | | | | | | | |
|---|---|---|---|---|---|---|---|---|
| V | 8969 | hCoV-19/Japan/PG-8712/2020\|EPI_ISL_686308\|2020-08 | ···················· | ···················· | ····················· | ···················· | ···················· | ···················· |
| V | 8970 | hCoV-19/Japan/PG-8713/2020\|EPI_ISL_686309\|2020-08 | ···················· | ···················· | ····················· | ···················· | ···················· | ···················· |
| V | 8971 | hCoV-19/Japan/PG-8714/2020\|EPI_ISL_686310\|2020-08 | ···················· | ···················· | ····················· | ···················· | ···················· | ···················· |
| V | 8972 | hCoV-19/Japan/PG-8715/2020\|EPI_ISL_686311\|2020-08 | ···················· | ···················· | ····················· | ···················· | ···················· | ···················· |
| V | 8973 | hCoV-19/Japan/PG-8716/2020\|EPI_ISL_686312\|2020-08 | ···················· | ···················· | ····················· | ···················· | ···················· | ···················· |
| V | 8974 | hCoV-19/Japan/PG-8717/2020\|EPI_ISL_686313\|2020-08 | ···················· | ···················· | ····················· | ···················· | ···················· | ···················· |
| V | 8975 | hCoV-19/Japan/PG-8718/2020\|EPI_ISL_686314\|2020-08 | ···················· | ···················· | ····················· | ···················· | ···················· | ···················· |
| V | 8976 | hCoV-19/Japan/PG-8719/2020\|EPI_ISL_686315\|2020-08 | ···················· | ···················· | ····················· | ···················· | ···················· | ···················· |
| V | 8977 | hCoV-19/Japan/PG-8720/2020\|EPI_ISL_686316\|2020-08 | ···················· | ···················· | ····················· | ···················· | ···················· | ···················· |
| V | 8978 | hCoV-19/Japan/PG-8721/2020\|EPI_ISL_686317\|2020-08 | ···················· | ···················· | ····················· | ···················· | ···················· | ···················· |
| V | 8979 | hCoV-19/Japan/PG-8722/2020\|EPI_ISL_686318\|2020-08 | ···················· | ···················· | ····················· | ···················· | ···················· | ···················· |
| V | 8980 | hCoV-19/Japan/PG-8723/2020\|EPI_ISL_686319\|2020-08 | ···················· | ···················· | ····················· | ···················· | ···················· | ···················· |
| V | 8981 | hCoV-19/Japan/PG-8724/2020\|EPI_ISL_686320\|2020-08 | ···················· | ···················· | ····················· | ···················· | ···················· | ···················· |
| V | 8982 | hCoV-19/Japan/PG-8725/2020\|EPI_ISL_686321\|2020-08 | ···················· | ···················· | ····················· | ···················· | ···················· | ···················· |
| V | 8983 | hCoV-19/Japan/PG-8726/2020\|EPI_ISL_686322\|2020-08 | ···················· | ···················· | ····················· | ···················· | ···················· | ···················· |
| V | 8984 | hCoV-19/Japan/PG-8728/2020\|EPI_ISL_686324\|2020-08 | ···················· | ···················· | ····················· | ···················· | ···················· | ···················· |
| V | 8985 | hCoV-19/Japan/PG-8501/2020\|EPI_ISL_687600\|2020-08 | ···················· | ···················· | ····················· | ···················· | ···················· | ···················· |
| V | 8986 | hCoV-19/Japan/PG-8502/2020\|EPI_ISL_687601\|2020-09 | ···················· | ···················· | ····················· | ···················· | ···················· | ···················· |
| V | 8987 | hCoV-19/Japan/PG-8503/2020\|EPI_ISL_687602\|2020-09 | ···················· | ···················· | ····················· | ···················· | ···················· | ···················· |
| V | 8988 | hCoV-19/Japan/PG-8504/2020\|EPI_ISL_687603\|2020-09 | ···················· | ···················· | ····················· | ···················· | ···················· | ···················· |
| V | 8989 | hCoV-19/Japan/PG-8505/2020\|EPI_ISL_687604\|2020-09 | ···················· | ···················· | ···T················· | ···················· | ···················· | ···················· |
| V | 8990 | hCoV-19/Japan/PG-8506/2020\|EPI_ISL_687605\|2020-09 | ···················· | ···················· | ····················· | ···················· | ···················· | ···················· |
| V | 8991 | hCoV-19/Japan/PG-8507/2020\|EPI_ISL_687606\|2020-09 | ···················· | ···················· | ····················· | ···················· | ···················· | ···················· |
| V | 8992 | hCoV-19/Japan/PG-8508/2020\|EPI_ISL_687607\|2020-09 | ···················· | ···················· | ····················· | ···················· | ···················· | ···················· |
| V | 8993 | hCoV-19/Japan/PG-8509/2020\|EPI_ISL_687608\|2020-09 | ···················· | ···················· | ····················· | ···················· | ···················· | ···················· |
| V | 8994 | hCoV-19/Japan/PG-8510/2020\|EPI_ISL_687609\|2020-07 | ···················· | ···················· | ····················· | ···················· | ···················· | ···················· |
| V | 8995 | hCoV-19/Japan/PG-8511/2020\|EPI_ISL_687610\|2020-07 | ···················· | ···················· | ····················· | ···················· | ···················· | ···················· |
| V | 8996 | hCoV-19/Japan/PG-8512/2020\|EPI_ISL_687611\|2020-07 | ···················· | ···················· | ····················· | ···················· | ···················· | ···················· |
| V | 8997 | hCoV-19/Japan/PG-7347/2020\|EPI_ISL_687614\|2020-08 | ···················· | ···················· | ····················· | ···················· | ···················· | ···················· |
| V | 8998 | hCoV-19/Japan/PG-6657/2020\|EPI_ISL_687616\|2020-07 | ···················· | ···················· | ····················· | ···················· | ···················· | ···················· |
| V | 8999 | hCoV-19/Japan/PG-6737/2020\|EPI_ISL_687617\|2020-08 | ···················· | ···················· | ····················· | ···················· | ···················· | ···················· |
| V | 9000 | hCoV-19/Japan/PG-6817/2020\|EPI_ISL_687618\|2020-04 | ···················· | ···················· | ····················· | ···················· | ···················· | ···················· |
| V | 9001 | hCoV-19/Japan/PG-7183/2020\|EPI_ISL_687619\|2020-07 | ···················· | ···················· | ····················· | ···················· | ···················· | ···················· |
| V | 9002 | hCoV-19/Japan/PG-7240/2020\|EPI_ISL_687620\|2020-08 | ···················· | ···················· | ····················· | ···················· | ···················· | ···················· |
| V | 9003 | hCoV-19/Japan/PG-6510/2020\|EPI_ISL_687624\|2020-07 | ···················· | ···················· | ····················· | ···················· | ···················· | ···················· |
| V | 9004 | hCoV-19/Japan/PG-6511/2020\|EPI_ISL_687625\|2020-07 | ···················· | ···················· | ····················· | ···················· | ···················· | ···················· |
| V | 9005 | hCoV-19/Japan/PG-6512/2020\|EPI_ISL_687626\|2020-07 | ···················· | ···················· | ····················· | ···················· | ···················· | ···················· |
| V | 9006 | hCoV-19/Japan/PG-6513/2020\|EPI_ISL_687627\|2020-07 | ···················· | ···················· | ····················· | ···················· | ···················· | ···················· |
| V | 9007 | hCoV-19/Japan/PG-6514/2020\|EPI_ISL_687628\|2020-07 | ···················· | ···················· | ····················· | ···················· | ···················· | ···················· |
| V | 9008 | hCoV-19/Japan/PG-6515/2020\|EPI_ISL_687629\|2020-07 | ···················· | ···················· | ····················· | ···················· | ···················· | ···················· |
| V | 9009 | hCoV-19/Japan/PG-6516/2020\|EPI_ISL_687630\|2020-07 | ···················· | ···················· | ····················· | ···················· | ···················· | ···················· |
| V | 9010 | hCoV-19/Japan/PG-6517/2020\|EPI_ISL_687631\|2020-07 | ···················· | ···················· | ····················· | ···················· | ···················· | ···················· |
| V | 9011 | hCoV-19/Japan/PG-6518/2020\|EPI_ISL_687632\|2020-07 | ···················· | ···················· | ····················· | ···················· | ···················· | ···················· |
| V | 9012 | hCoV-19/Japan/PG-6519/2020\|EPI_ISL_687633\|2020-07 | ···················· | ···················· | ····················· | ···················· | ···················· | ···················· |
| V | 9013 | hCoV-19/Japan/PG-6520/2020\|EPI_ISL_687634\|2020-07 | ···················· | ···················· | ····················· | ···················· | ···················· | ···················· |
| V | 9014 | hCoV-19/Japan/PG-6521/2020\|EPI_ISL_687635\|2020-07 | ···················· | ···················· | ····················· | ···················· | ···················· | ···················· |
| V | 9015 | hCoV-19/Japan/PG-6522/2020\|EPI_ISL_687636\|2020-07 | ···················· | ···················· | ····················· | ···················· | ···················· | ···················· |
| V | 9016 | hCoV-19/Japan/PG-6523/2020\|EPI_ISL_687637\|2020-07 | ···················· | ···················· | ····················· | ···················· | ···················· | ···················· |
| V | 9017 | hCoV-19/Japan/PG-6524/2020\|EPI_ISL_687638\|2020-07 | ···················· | ···················· | ····················· | ···················· | ···················· | ···················· |
| V | 9018 | hCoV-19/Japan/PG-6525/2020\|EPI_ISL_687639\|2020-07 | ···················· | ···················· | ····················· | ···················· | ···················· | ···················· |
| V | 9019 | hCoV-19/Japan/PG-6526/2020\|EPI_ISL_687640\|2020-07 | ···················· | ···················· | ····················· | ···················· | ···················· | ···················· |
| V | 9020 | hCoV-19/Japan/PG-6527/2020\|EPI_ISL_687641\|2020-07 | ···················· | ···················· | ····················· | ···················· | ···················· | ···················· |
| V | 9021 | hCoV-19/Japan/PG-6528/2020\|EPI_ISL_687642\|2020-08 | ···················· | ···················· | ····················· | ···················· | ···················· | ···················· |
| V | 9022 | hCoV-19/Japan/PG-6529/2020\|EPI_ISL_687643\|2020-08 | ···················· | ···················· | ····················· | ···················· | ···················· | ···················· |
| V | 9023 | hCoV-19/Japan/PG-6530/2020\|EPI_ISL_687644\|2020-08 | ···················· | ···················· | ····················· | ···················· | ···················· | ···················· |
| V | 9024 | hCoV-19/Japan/PG-6531/2020\|EPI_ISL_687645\|2020-08 | ···················· | ···················· | ····················· | ···················· | ···················· | ···················· |
| V | 9025 | hCoV-19/Japan/PG-6532/2020\|EPI_ISL_687646\|2020-08 | ···················· | ···················· | ····················· | ···················· | ···················· | ···················· |
| V | 9026 | hCoV-19/Japan/PG-6533/2020\|EPI_ISL_687647\|2020-08 | ···················· | ···················· | ····················· | ···················· | ···················· | ···················· |
| V | 9027 | hCoV-19/Japan/PG-6534/2020\|EPI_ISL_687648\|2020-08 | ···················· | ···················· | ····················· | ···················· | ···················· | ···················· |
| V | 9028 | hCoV-19/Japan/PG-6535/2020\|EPI_ISL_687649\|2020-08 | ···················· | ···················· | ····················· | ···················· | ···················· | ···················· |
| V | 9029 | hCoV-19/Japan/PG-6536/2020\|EPI_ISL_687650\|2020-08 | ···················· | ···················· | ····················· | ···················· | ···················· | ···················· |
| V | 9030 | hCoV-19/Japan/PG-6537/2020\|EPI_ISL_687651\|2020-08 | ···················· | ···················· | ····················· | ···················· | ···················· | ···················· |
| V | 9031 | hCoV-19/Japan/PG-6538/2020\|EPI_ISL_687652\|2020-08 | ···················· | ···················· | ····················· | ···················· | ···················· | ···················· |
| V | 9032 | hCoV-19/Japan/PG-6539/2020\|EPI_ISL_687653\|2020-08 | ···················· | ···················· | ····················· | ···················· | ···················· | ···················· |
| V | 9033 | hCoV-19/Japan/PG-6540/2020\|EPI_ISL_687654\|2020-08 | ···················· | ···················· | ····················· | ···················· | ···················· | ···················· |
| V | 9034 | hCoV-19/Japan/PG-5812/2020\|EPI_ISL_688929\|2020-08 | ···················· | ···················· | ····················· | ···················· | ···················· | ···················· |
| V | 9035 | hCoV-19/Japan/PG-5813/2020\|EPI_ISL_688930\|2020-08 | ···················· | ···················· | ····················· | ···················· | ···················· | ···················· |
| V | 9036 | hCoV-19/Japan/PG-5814/2020\|EPI_ISL_688931\|2020-08 | ···················· | ···················· | ····················· | ···················· | ···················· | ···················· |
| V | 9037 | hCoV-19/Japan/PG-5815/2020\|EPI_ISL_688932\|2020-08 | ···················· | ···················· | ····················· | ···················· | ···················· | ···················· |
| V | 9038 | hCoV-19/Japan/PG-5816/2020\|EPI_ISL_688933\|2020-08 | ···················· | ···················· | ····················· | ···················· | ···················· | ···················· |
| V | 9039 | hCoV-19/Japan/PG-5817/2020\|EPI_ISL_688934\|2020-08 | ···················· | ···················· | ····················· | ···················· | ···················· | ···················· |
| V | 9040 | hCoV-19/Japan/PG-5818/2020\|EPI_ISL_688935\|2020-08 | ···················· | ···················· | ····················· | ···················· | ···················· | ···················· |
| V | 9041 | hCoV-19/Japan/PG-5819/2020\|EPI_ISL_688936\|2020-08 | ···················· | ···················· | ····················· | ···················· | ···················· | ···················· |
| V | 9042 | hCoV-19/Japan/PG-5820/2020\|EPI_ISL_688937\|2020-08 | ···················· | ···················· | ····················· | ···················· | ···················· | ···················· |
| V | 9043 | hCoV-19/Japan/PG-5821/2020\|EPI_ISL_688938\|2020-08 | ···················· | ···················· | ····················· | ···················· | ···················· | ···················· |
| V | 9044 | hCoV-19/Japan/PG-5822/2020\|EPI_ISL_688939\|2020-08 | ···················· | ···················· | ····················· | ···················· | ···················· | ···················· |
| V | 9045 | hCoV-19/Japan/PG-5823/2020\|EPI_ISL_688940\|2020-08 | ···················· | ···················· | ····················· | ···················· | ···················· | ···················· |
| V | 9046 | hCoV-19/Japan/PG-5824/2020\|EPI_ISL_688941\|2020-08 | ···················· | ···················· | ····················· | ···················· | ···················· | ···················· |
| V | 9047 | hCoV-19/Japan/PG-5825/2020\|EPI_ISL_688942\|2020-08 | ···················· | ···················· | ····················· | ···················· | ···················· | ···················· |
| V | 9048 | hCoV-19/Japan/PG-5826/2020\|EPI_ISL_688943\|2020-08 | ···················· | ···················· | ····················· | ···················· | ···················· | ···················· |

| | | | | | | | |
|---|---|---|---|---|---|---|---|
| V | 9049 hCoV-19/Japan/PG-5827/2020\|EPI_ISL_688944\|2020-08 | .................... | .................... | .................... | .................... | .................... | .................... |
| V | 9050 hCoV-19/Japan/PG-5828/2020\|EPI_ISL_688945\|2020-08 | .................... | .................... | .................... | .................... | .................... | .................... |
| V | 9051 hCoV-19/Japan/PG-5829/2020\|EPI_ISL_688946\|2020-08 | .................... | .................... | .................... | .................... | .................... | .................... |
| V | 9052 hCoV-19/Japan/PG-5830/2020\|EPI_ISL_688947\|2020-08 | .................... | .................... | .................... | .................... | .................... | .................... |
| V | 9053 hCoV-19/Japan/PG-5831/2020\|EPI_ISL_688948\|2020-08 | .................... | .................... | .................... | .................... | .................... | .................... |
| V | 9054 hCoV-19/Japan/PG-5832/2020\|EPI_ISL_688949\|2020-08 | .................... | .................... | .................... | .................... | .................... | .................... |
| V | 9055 hCoV-19/Japan/PG-5833/2020\|EPI_ISL_688950\|2020-08 | .................... | .................... | .................... | .................... | .................... | .................... |
| V | 9056 hCoV-19/Japan/PG-5834/2020\|EPI_ISL_688951\|2020-08 | .................... | .................... | .................... | .................... | .................... | .................... |
| V | 9057 hCoV-19/Japan/PG-5835/2020\|EPI_ISL_688952\|2020-08 | .................... | .................... | .................... | .................... | .................... | .................... |
| V | 9058 hCoV-19/Japan/PG-5836/2020\|EPI_ISL_688953\|2020-08 | .................... | .................... | .................... | .................... | .................... | .................... |
| V | 9059 hCoV-19/Japan/PG-5837/2020\|EPI_ISL_688954\|2020-08 | .................... | .................... | .................... | .................... | .................... | .................... |
| V | 9060 hCoV-19/Japan/PG-5838/2020\|EPI_ISL_688955\|2020-08 | .................... | .................... | .................... | .................... | .................... | .................... |
| V | 9061 hCoV-19/Japan/PG-5839/2020\|EPI_ISL_688956\|2020-08 | .................... | .................... | .................... | .................... | .................... | .................... |
| V | 9062 hCoV-19/Japan/PG-5840/2020\|EPI_ISL_688957\|2020-08 | .................... | .................... | .................... | .................... | .................... | .................... |
| V | 9063 hCoV-19/Japan/PG-5841/2020\|EPI_ISL_688958\|2020-08 | .................... | .................... | .................... | .................... | .................... | .................... |
| V | 9064 hCoV-19/Japan/PG-5842/2020\|EPI_ISL_688959\|2020-08 | .................... | .................... | .................... | .................... | .................... | .................... |
| V | 9065 hCoV-19/Japan/PG-5843/2020\|EPI_ISL_688960\|2020-08 | .................... | .................... | .................... | .................... | .................... | .................... |
| V | 9066 hCoV-19/Japan/PG-5844/2020\|EPI_ISL_688961\|2020-08 | .................... | .................... | .................... | .................... | .................... | .................... |
| V | 9067 hCoV-19/Japan/PG-5845/2020\|EPI_ISL_688962\|2020-08 | .................... | .................... | .................... | .................... | .................... | .................... |
| V | 9068 hCoV-19/Japan/PG-5846/2020\|EPI_ISL_688963\|2020-08 | .................... | .................... | .................... | .................... | .................... | .................... |
| V | 9069 hCoV-19/Japan/PG-5847/2020\|EPI_ISL_688964\|2020-08 | .................... | .................... | .................... | .................... | .................... | .................... |
| V | 9070 hCoV-19/Japan/PG-5848/2020\|EPI_ISL_688965\|2020-08 | .................... | .................... | .................... | .................... | .................... | .................... |
| V | 9071 hCoV-19/Japan/PG-5849/2020\|EPI_ISL_688966\|2020-08 | .................... | .................... | .................... | .................... | .................... | .................... |
| V | 9072 hCoV-19/Japan/PG-5850/2020\|EPI_ISL_688967\|2020-08 | .................... | .................... | .................... | .................... | .................... | .................... |
| V | 9073 hCoV-19/Japan/PG-5851/2020\|EPI_ISL_688968\|2020-08 | .................... | .................... | .................... | .................... | .................... | .................... |
| V | 9074 hCoV-19/Japan/PG-5852/2020\|EPI_ISL_688969\|2020-08 | .................... | .................... | .................... | .................... | .................... | .................... |
| V | 9075 hCoV-19/Japan/PG-5853/2020\|EPI_ISL_688970\|2020-08 | .................... | .................... | .................... | .................... | .................... | .................... |
| V | 9076 hCoV-19/Japan/PG-5854/2020\|EPI_ISL_688971\|2020-08 | .................... | .................... | .................... | .................... | .................... | .................... |
| V | 9077 hCoV-19/Japan/PG-5855/2020\|EPI_ISL_688972\|2020-08 | .................... | .................... | .................... | .................... | .................... | .................... |
| V | 9078 hCoV-19/Japan/PG-5856/2020\|EPI_ISL_688973\|2020-08 | .................... | .................... | .................... | .................... | .................... | .................... |
| V | 9079 hCoV-19/Japan/PG-5857/2020\|EPI_ISL_688974\|2020-08 | .................... | .................... | .................... | .................... | .................... | .................... |
| V | 9080 hCoV-19/Japan/PG-5858/2020\|EPI_ISL_688975\|2020-08 | .................... | .................... | .................... | .................... | .................... | .................... |
| V | 9081 hCoV-19/Japan/PG-5859/2020\|EPI_ISL_688976\|2020-08 | .................... | .................... | .................... | .................... | .................... | .................... |
| V | 9082 hCoV-19/Japan/PG-5860/2020\|EPI_ISL_688977\|2020-08 | .................... | .................... | .................... | .................... | .................... | .................... |
| V | 9083 hCoV-19/Japan/PG-5861/2020\|EPI_ISL_688978\|2020-08 | .................... | .................... | .................... | .................... | .................... | .................... |
| V | 9084 hCoV-19/Japan/PG-5862/2020\|EPI_ISL_688979\|2020-08 | .................... | .................... | .................... | .................... | .................... | .................... |
| V | 9085 hCoV-19/Japan/PG-5863/2020\|EPI_ISL_688980\|2020-08 | .................... | .................... | .................... | .................... | .................... | .................... |
| V | 9086 hCoV-19/Japan/PG-5866/2020\|EPI_ISL_688981\|2020-08 | .................... | .................... | .................... | .................... | .................... | .................... |
| V | 9087 hCoV-19/Japan/PG-5867/2020\|EPI_ISL_688982\|2020-08 | .................... | .................... | .................... | .................... | .................... | .................... |
| V | 9088 hCoV-19/Japan/PG-5868/2020\|EPI_ISL_688983\|2020-08 | .................... | .................... | .................... | .................... | .................... | .................... |
| V | 9089 hCoV-19/Japan/PG-5869/2020\|EPI_ISL_688984\|2020-08 | .................... | .................... | .................... | .................... | .................... | .................... |
| V | 9090 hCoV-19/Japan/PG-5870/2020\|EPI_ISL_688985\|2020-08 | .................... | .................... | .................... | .................... | .................... | .................... |
| V | 9091 hCoV-19/Japan/PG-3849/2020\|EPI_ISL_692100\|2020-07 | .................... | .................... | .................... | .................... | .................... | .................... |
| V | 9092 hCoV-19/Japan/PG-3850/2020\|EPI_ISL_692101\|2020-07 | .................... | .................... | .................... | .................... | .................... | .................... |
| V | 9093 hCoV-19/Japan/PG-3851/2020\|EPI_ISL_692102\|2020-07 | .................... | .................... | .................... | .................... | .................... | .................... |
| V | 9094 hCoV-19/Japan/PG-3852/2020\|EPI_ISL_692103\|2020-07 | .................... | .................... | .................... | .................... | .................... | .................... |
| V | 9095 hCoV-19/Japan/PG-3853/2020\|EPI_ISL_692104\|2020-07 | .................... | .................... | .................... | .................... | .................... | .................... |
| V | 9096 hCoV-19/Japan/PG-3854/2020\|EPI_ISL_692105\|2020-07 | .................... | .................... | .................... | .................... | .................... | .................... |
| V | 9097 hCoV-19/Japan/PG-3855/2020\|EPI_ISL_692106\|2020-07 | .................... | .................... | .................... | .................... | .................... | .................... |
| V | 9098 hCoV-19/Japan/PG-3856/2020\|EPI_ISL_692107\|2020-07 | .................... | .................... | .................... | .................... | .................... | .................... |
| V | 9099 hCoV-19/Japan/PG-3857/2020\|EPI_ISL_692108\|2020-07 | .................... | .................... | .................... | .................... | .................... | .................... |
| V | 9100 hCoV-19/Japan/PG-3858/2020\|EPI_ISL_692109\|2020-07 | .................... | .................... | .................... | .................... | .................... | .................... |
| V | 9101 hCoV-19/Japan/PG-3859/2020\|EPI_ISL_692110\|2020-07 | .................... | .................... | .................... | .................... | .................... | .................... |
| V | 9102 hCoV-19/Japan/PG-3860/2020\|EPI_ISL_692111\|2020-07 | .................... | .................... | .................... | .................... | .................... | .................... |
| V | 9103 hCoV-19/Japan/PG-3861/2020\|EPI_ISL_692112\|2020-07 | .................... | .................... | .................... | .................... | .................... | .................... |
| V | 9104 hCoV-19/Japan/PG-3862/2020\|EPI_ISL_692113\|2020-07 | .................... | .................... | .................... | .................... | .................... | .................... |
| V | 9105 hCoV-19/Japan/PG-3863/2020\|EPI_ISL_692114\|2020-07 | .................... | .................... | .................... | .................... | .................... | .................... |
| V | 9106 hCoV-19/Japan/PG-3864/2020\|EPI_ISL_692115\|2020-07 | .................... | .................... | .................... | .................... | .................... | .................... |
| V | 9107 hCoV-19/Japan/PG-3865/2020\|EPI_ISL_692116\|2020-07 | .................... | .................... | .................... | .................... | .................... | .................... |
| V | 9108 hCoV-19/Japan/PG-3866/2020\|EPI_ISL_692117\|2020-07 | .................... | .................... | .................... | .................... | .................... | .................... |
| V | 9109 hCoV-19/Japan/PG-3867/2020\|EPI_ISL_692118\|2020-07 | .................... | .................... | .................... | .................... | .................... | .................... |
| V | 9110 hCoV-19/Japan/PG-3868/2020\|EPI_ISL_692119\|2020-06 | .................... | .................... | .................... | .................... | .................... | .................... |
| V | 9111 hCoV-19/Japan/PG-3869/2020\|EPI_ISL_692120\|2020-06 | .................... | .................... | .................... | .................... | .................... | .................... |
| V | 9112 hCoV-19/Japan/PG-3870/2020\|EPI_ISL_692121\|2020-06 | .................... | .................... | .................... | .................... | .................... | .................... |
| V | 9113 hCoV-19/Japan/PG-3871/2020\|EPI_ISL_692122\|2020-06 | .................... | .................... | .................... | .................... | .................... | .................... |
| V | 9114 hCoV-19/Japan/PG-3872/2020\|EPI_ISL_692123\|2020-06 | .................... | .................... | .................... | .................... | .................... | .................... |
| V | 9115 hCoV-19/Japan/PG-3873/2020\|EPI_ISL_692124\|2020-06 | .................... | .................... | .................... | .................... | .................... | .................... |
| V | 9117 hCoV-19/Japan/PG-3875/2020\|EPI_ISL_692126\|2020-06 | .................... | .................... | .................... | .................... | .................... | .................... |
| V | 9118 hCoV-19/Japan/PG-3876/2020\|EPI_ISL_692127\|2020-06 | .................... | .................... | .................... | .................... | .................... | .................... |
| V | 9119 hCoV-19/Japan/PG-3877/2020\|EPI_ISL_692128\|2020-06 | .................... | .................... | .................... | .................... | .................... | .................... |
| V | 9120 hCoV-19/Japan/PG-3878/2020\|EPI_ISL_692129\|2020-07 | .................... | .................... | .................... | .................... | .................... | .................... |
| V | 9121 hCoV-19/Japan/PG-3879/2020\|EPI_ISL_692130\|2020-07 | .................... | .................... | .................... | .................... | .................... | .................... |
| V | 9122 hCoV-19/Japan/PG-3880/2020\|EPI_ISL_692131\|2020-07 | .................... | .................... | .................... | .................... | .................... | .................... |
| V | 9123 hCoV-19/Japan/PG-3881/2020\|EPI_ISL_692132\|2020-07 | .................... | .................... | .................... | .................... | .................... | .................... |
| V | 9124 hCoV-19/Japan/PG-3882/2020\|EPI_ISL_692133\|2020-07 | .................... | .................... | .................... | .................... | .................... | .................... |
| V | 9125 hCoV-19/Japan/PG-3883/2020\|EPI_ISL_692134\|2020-07 | .................... | .................... | .................... | .................... | .................... | .................... |
| V | 9126 hCoV-19/Japan/PG-3884/2020\|EPI_ISL_692135\|2020-07 | .................... | .A.................. | .................... | .................... | .................... | .................... |
| V | 9127 hCoV-19/Japan/PG-3885/2020\|EPI_ISL_692136\|2020-07 | .................... | .A.................. | .................... | .................... | .................... | .................... |
| V | 9128 hCoV-19/Japan/PG-3886/2020\|EPI_ISL_692137\|2020-07 | .................... | .................... | .................... | .................... | .................... | .................... |
| V | 9129 hCoV-19/Japan/PG-3887/2020\|EPI_ISL_692138\|2020-07 | .................... | .................... | .................... | .................... | .................... | .................... |

| | | | | | | | | |
|---|---|---|---|---|---|---|---|---|
| V | 9130 | hCoV-19/Japan/PG-3888/2020\|EPI_ISL_692139\|2020-07 | ···················· | ···················· | ···················· | ···················· | ···················· | ···················· |
| V | 9131 | hCoV-19/Japan/PG-3889/2020\|EPI_ISL_692140\|2020-07 | ···················· | ···················· | ···················· | ···················· | ···················· | ···················· |
| V | 9132 | hCoV-19/Japan/PG-3890/2020\|EPI_ISL_692141\|2020-07 | ···················· | ···················· | ···················· | ···················· | ···················· | ···················· |
| V | 9133 | hCoV-19/Japan/PG-3891/2020\|EPI_ISL_692142\|2020-07 | ···················· | ···················· | ···················· | ···················· | ···················· | ···················· |
| V | 9134 | hCoV-19/Japan/PG-3892/2020\|EPI_ISL_692143\|2020-07 | ···················· | ···················· | ···················· | ···················· | ···················· | ···················· |
| V | 9135 | hCoV-19/Japan/PG-3893/2020\|EPI_ISL_692144\|2020-07 | ···················· | ···················· | ···················· | ···················· | ···················· | ···················· |
| V | 9136 | hCoV-19/Japan/PG-3894/2020\|EPI_ISL_692145\|2020-07 | ···················· | ···················· | ···················· | ···················· | ···················· | ···················· |
| V | 9137 | hCoV-19/Japan/PG-3895/2020\|EPI_ISL_692146\|2020-07 | ···················· | ···················· | ···················· | ···················· | ···················· | ···················· |
| V | 9138 | hCoV-19/Japan/PG-3896/2020\|EPI_ISL_692147\|2020-07 | ···················· | ···················· | ···················· | ···················· | ···················· | ···················· |
| V | 9139 | hCoV-19/Japan/PG-3897/2020\|EPI_ISL_692148\|2020-07 | ···················· | ···················· | ···················· | ···················· | ···················· | ···················· |
| V | 9140 | hCoV-19/Japan/PG-3898/2020\|EPI_ISL_692149\|2020-07 | ···················· | ···················· | ···················· | ···················· | ···················· | ···················· |
| V | 9141 | hCoV-19/Japan/PG-3899/2020\|EPI_ISL_692150\|2020-07 | ···················· | ···················· | ···················· | ···················· | ···················· | ···················· |
| V | 9142 | hCoV-19/Japan/PG-3900/2020\|EPI_ISL_692151\|2020-07 | ···················· | ···················· | ···················· | ···················· | ···················· | ···················· |
| V | 9143 | hCoV-19/Japan/PG-3901/2020\|EPI_ISL_692152\|2020-07 | ···················· | ···················· | ···················· | ···················· | ···················· | ···················· |
| V | 9144 | hCoV-19/Japan/PG-3902/2020\|EPI_ISL_692153\|2020-07 | ···················· | ···················· | ···················· | ···················· | ···················· | ···················· |
| V | 9146 | hCoV-19/Japan/PG-3904/2020\|EPI_ISL_692155\|2020-07 | ···················· | ···················· | ···················· | ···················· | ···················· | ···················· |
| V | 9147 | hCoV-19/Japan/PG-3905/2020\|EPI_ISL_692156\|2020-07 | ···················· | ···················· | ···················· | ···················· | ···················· | ···················· |
| V | 9148 | hCoV-19/Japan/PG-3906/2020\|EPI_ISL_692157\|2020-07 | ···················· | ···················· | ···················· | ···················· | ···················· | ···················· |
| V | 9149 | hCoV-19/Japan/PG-3907/2020\|EPI_ISL_692158\|2020-07 | ···················· | ···················· | ···················· | ···················· | ···················· | ···················· |
| V | 9150 | hCoV-19/Japan/PG-3908/2020\|EPI_ISL_692159\|2020-07 | ···················· | ···················· | ···················· | ···················· | ···················· | ···················· |
| V | 9151 | hCoV-19/Japan/PG-3909/2020\|EPI_ISL_692160\|2020-07 | ···················· | ···················· | ···················· | ···················· | ···················· | ···················· |
| V | 9152 | hCoV-19/Japan/PG-3910/2020\|EPI_ISL_692161\|2020-07 | ···················· | ···················· | ···················· | ···················· | ···················· | ···················· |
| V | 9153 | hCoV-19/Japan/PG-3911/2020\|EPI_ISL_692162\|2020-06 | ···················· | ···················· | ···················· | ···················· | ···················· | ···················· |
| V | 9154 | hCoV-19/Japan/PG-3912/2020\|EPI_ISL_692163\|2020-06 | ···················· | ···················· | ···················· | ···················· | ···················· | ···················· |
| V | 9155 | hCoV-19/Japan/PG-3913/2020\|EPI_ISL_692164\|2020-07 | ···················· | ···················· | ···················· | ···················· | ···················· | ···················· |
| V | 9156 | hCoV-19/Japan/PG-3914/2020\|EPI_ISL_692165\|2020-06 | ···················· | ···················· | ···················· | ···················· | ···················· | ···················· |
| V | 9157 | hCoV-19/England/204683210/2020\|EPI_ISL_693431\|2020 | ···················· | ···················· | ···················· | ···················· | ·····T·············· | ···················· |
| V | 9158 | hCoV-19/England/204691017/2020\|EPI_ISL_693432\|2020 | ···················· | ···················· | ···················· | ···················· | ···················· | ···················· |
| V | 9159 | hCoV-19/England/204691035/2020\|EPI_ISL_693433\|2020 | ···················· | ···················· | ···················· | ···················· | ···················· | ···················· |
| V | 9160 | hCoV-19/England/204691055/2020\|EPI_ISL_693434\|2020 | ···················· | ···················· | ···················· | ···················· | ···················· | ···················· |
| V | 9161 | hCoV-19/England/204691061/2020\|EPI_ISL_693435\|2020 | ···················· | ···················· | ···················· | ···················· | ···················· | ···················· |
| V | 9162 | hCoV-19/England/204691065/2020\|EPI_ISL_693436\|2020 | ···················· | ···················· | ···················· | ···················· | ···················· | ···················· |
| V | 9163 | hCoV-19/England/204691072/2020\|EPI_ISL_693437\|2020 | ···················· | ···················· | ···················· | ···················· | ···················· | ···················· |
| V | 9164 | hCoV-19/England/204691122/2020\|EPI_ISL_693438\|2020 | ···················· | ···················· | ···················· | ···················· | ···················· | ···················· |
| V | 9165 | hCoV-19/England/204691129/2020\|EPI_ISL_693439\|2020 | ···················· | ···················· | ···················· | ···················· | ···················· | ···················· |
| V | 9166 | hCoV-19/England/204691195/2020\|EPI_ISL_693440\|2020 | ···················· | ···················· | ···················· | ···················· | ·····T·············· | ···················· |
| V | 9167 | hCoV-19/England/204691197/2020\|EPI_ISL_693441\|2020 | ···················· | ···················· | ···················· | ···················· | ···················· | ···················· |
| V | 9168 | hCoV-19/England/204691822/2020\|EPI_ISL_693442\|2020 | ···················· | ···················· | ···················· | ···················· | ···················· | ···················· |
| V | 9169 | hCoV-19/England/204691824/2020\|EPI_ISL_693443\|2020 | ···················· | ···················· | ···················· | ···················· | ···················· | ···················· |
| V | 9170 | hCoV-19/England/204691826/2020\|EPI_ISL_693444\|2020 | ···················· | ···················· | ···················· | ···················· | ···················· | ···················· |
| V | 9171 | hCoV-19/England/204691900/2020\|EPI_ISL_693445\|2020 | ···················· | ···················· | ···················· | ···················· | ···················· | ···················· |
| V | 9172 | hCoV-19/England/204691901/2020\|EPI_ISL_693446\|2020 | ···················· | ···················· | ···················· | ···················· | ···················· | ···················· |
| V | 9173 | hCoV-19/England/204691902/2020\|EPI_ISL_693447\|2020 | ···················· | ···················· | ···················· | ···················· | ·····T·············· | ···················· |
| V | 9174 | hCoV-19/England/204691916/2020\|EPI_ISL_693448\|2020 | ···················· | ···················· | ···················· | ···················· | ···················· | ···················· |
| V | 9175 | hCoV-19/England/204691940/2020\|EPI_ISL_693449\|2020 | ···················· | ···················· | ···················· | ···················· | ···················· | ···················· |
| V | 9176 | hCoV-19/England/204691942/2020\|EPI_ISL_693450\|2020 | ···················· | ···················· | ···················· | ···················· | ···················· | ···················· |
| V | 9177 | hCoV-19/England/204691943/2020\|EPI_ISL_693451\|2020 | ···················· | ···················· | ···················· | ···················· | ···················· | ···················· |
| V | 9178 | hCoV-19/England/204691946/2020\|EPI_ISL_693452\|2020 | ···················· | ···················· | ···················· | ···················· | ···················· | ···················· |
| V | 9179 | hCoV-19/England/204691948/2020\|EPI_ISL_693453\|2020 | ···················· | ···················· | ···················· | ···················· | ···················· | ···················· |
| V | 9180 | hCoV-19/England/204691949/2020\|EPI_ISL_693454\|2020 | ···················· | ···················· | ···················· | ···················· | ···················· | ···················· |
| V | 9181 | hCoV-19/England/204700011/2020\|EPI_ISL_693455\|2020 | ···················· | ···················· | ···················· | ···················· | ···················· | ···················· |
| V | 9182 | hCoV-19/England/204700012/2020\|EPI_ISL_693456\|2020 | ···················· | ···················· | ···················· | ···················· | ···················· | ···················· |
| V | 9183 | hCoV-19/England/204700064/2020\|EPI_ISL_693457\|2020 | ···················· | ···················· | ···················· | ···················· | ···················· | ···················· |
| V | 9184 | hCoV-19/England/204700071/2020\|EPI_ISL_693458\|2020 | ···················· | ···················· | ···················· | ···················· | ·····T·············· | ···················· |
| V | 9185 | hCoV-19/England/204700075/2020\|EPI_ISL_693459\|2020 | ···················· | ···················· | ···················· | ···················· | ·····T·············· | ···················· |
| V | 9186 | hCoV-19/England/204721173/2020\|EPI_ISL_693460\|2020 | ···················· | ···················· | ···················· | ···················· | ···················· | ···················· |
| V | 9187 | hCoV-19/England/204722566/2020\|EPI_ISL_693461\|2020 | ·······Y············ | ···················· | ···················· | ···················· | ···················· | ···················· |
| V | 9188 | hCoV-19/England/204722726/2020\|EPI_ISL_693462\|2020 | ···················· | ···················· | ···················· | ···················· | ···················· | ···················· |
| V | 9189 | hCoV-19/England/204740421/2020\|EPI_ISL_693463\|2020 | ···················· | ···················· | ···················· | ···················· | ·····T·············· | ···················· |
| V | 9190 | hCoV-19/England/204740422/2020\|EPI_ISL_693464\|2020 | ···················· | ···················· | ···················· | ···················· | ···················· | ···················· |
| V | 9191 | hCoV-19/England/204740425/2020\|EPI_ISL_693465\|2020 | ···················· | ···················· | ···················· | ···················· | ···················· | ···················· |
| V | 9192 | hCoV-19/England/204740428/2020\|EPI_ISL_693466\|2020 | ···················· | ···················· | ···················· | ···················· | ···················· | ···················· |
| V | 9193 | hCoV-19/England/204740527/2020\|EPI_ISL_693467\|2020 | ···················· | ···················· | ···················· | ···················· | ···················· | ···················· |
| V | 9194 | hCoV-19/England/204740528/2020\|EPI_ISL_693468\|2020 | ···················· | ···················· | ···················· | ···················· | ···················· | ···················· |
| V | 9195 | hCoV-19/England/204740531/2020\|EPI_ISL_693469\|2020 | ···················· | ···················· | ···················· | ···················· | ···················· | ···················· |
| V | 9196 | hCoV-19/England/204740539/2020\|EPI_ISL_693470\|2020 | ···················· | ···················· | ···················· | ···················· | ···················· | ···················· |
| V | 9197 | hCoV-19/Papua New Guinea/1/2020\|EPI_ISL_693471\|202 | ···················· | ···················· | ···················· | ···················· | ···················· | ···················· |
| V | 9198 | hCoV-19/Papua New Guinea/2/2020\|EPI_ISL_693472\|202 | ···················· | ···················· | ···················· | ···················· | ···················· | ···················· |
| V | 9199 | hCoV-19/Papua New Guinea/3/2020\|EPI_ISL_693473\|202 | ···················· | ···················· | ···················· | ···················· | ···················· | ···················· |
| V | 9200 | hCoV-19/Papua New Guinea/4/2020\|EPI_ISL_693474\|202 | ···················· | ···················· | ···················· | ···················· | ···················· | ···················· |
| V | 9201 | hCoV-19/Papua New Guinea/5/2020\|EPI_ISL_693475\|202 | ···················· | ···················· | ···················· | ···················· | ···················· | ···················· |
| V | 9202 | hCoV-19/Papua New Guinea/6/2020\|EPI_ISL_693476\|202 | ···················· | ···················· | ···················· | ···················· | ···················· | ···················· |
| V | 9203 | hCoV-19/Papua New Guinea/7/2020\|EPI_ISL_693477\|202 | ···················· | ···················· | ···················· | ···················· | ···················· | ···················· |
| V | 9204 | hCoV-19/Papua New Guinea/8/2020\|EPI_ISL_693478\|202 | ···················· | ···················· | ···················· | ···················· | ···················· | ···················· |
| V | 9205 | hCoV-19/Papua New Guinea/9/2020\|EPI_ISL_693479\|202 | ···················· | ···················· | ···················· | ···················· | ···················· | ···················· |
| V | 9206 | hCoV-19/Papua New Guinea/10/2020\|EPI_ISL_693480\|20 | ···················· | ···················· | ···················· | ···················· | ···················· | ···················· |
| V | 9207 | hCoV-19/Papua New Guinea/11/2020\|EPI_ISL_693481\|20 | ···················· | ···················· | ···················· | ···················· | ···················· | ···················· |
| V | 9208 | hCoV-19/Papua New Guinea/12/2020\|EPI_ISL_693482\|20 | ···················· | ···················· | ···················· | ···················· | ···················· | ···················· |
| V | 9209 | hCoV-19/Papua New Guinea/13/2020\|EPI_ISL_693483\|20 | ···················· | ···················· | ···················· | ···················· | ···················· | ···················· |
| V | 9210 | hCoV-19/France/ARA-70122/2020\|EPI_ISL_693490\|2020- | ···················· | ···················· | ···················· | ···················· | ···················· | ···················· |

| | | | | | | | | |
|---|---|---|---|---|---|---|---|---|
| V | 9211 | hCoV-19/France/ARA-70764/2020\|EPI_ISL_693491\|2020- | .................... | .................... | ..................... | .................... | .................... | .................... |
| V | 9212 | hCoV-19/France/ARA-7096/2020\|EPI_ISL_693492\|2020-0 | .................... | .................... | ..................... | .................... | .................... | .................... |
| V | 9213 | hCoV-19/France/ARA-71474/2020\|EPI_ISL_693493\|2020- | .................... | .................... | ..................... | .................... | .................... | .................... |
| V | 9214 | hCoV-19/France/ARA-71481/2020\|EPI_ISL_693494\|2020- | .................... | .................... | ..................... | .................... | .................... | .................... |
| V | 9215 | hCoV-19/France/ARA-71554/2020\|EPI_ISL_693495\|2020- | .................... | .................... | ..................... | .................... | .................... | .................... |
| V | 9216 | hCoV-19/France/ARA-72054/2020\|EPI_ISL_693496\|2020- | .................... | .................... | ..................... | .................... | .................... | .................... |
| V | 9217 | hCoV-19/Japan/PG-8619/2020\|EPI_ISL_686215\|2020-08 | .................... | .................... | ..................... | .................... | .................... | .................... |
| V | 9218 | hCoV-19/Japan/PG-8620/2020\|EPI_ISL_686216\|2020-08 | .................... | .................... | ..................... | .................... | .................... | .................... |
| V | 9219 | hCoV-19/Japan/PG-8621/2020\|EPI_ISL_686217\|2020-09 | .................... | .................... | ..................... | .................... | .................... | .................... |
| V | 9220 | hCoV-19/Japan/PG-8622/2020\|EPI_ISL_686218\|2020-09 | .................... | .................... | ..................... | .................... | .................... | .................... |
| V | 9221 | hCoV-19/Japan/PG-8623/2020\|EPI_ISL_686219\|2020-09 | .................... | .................... | ..................... | .................... | .................... | .................... |
| V | 9222 | hCoV-19/Japan/PG-8624/2020\|EPI_ISL_686220\|2020-09 | .................... | .................... | ..................... | .................... | .................... | .................... |
| V | 9223 | hCoV-19/Japan/PG-8625/2020\|EPI_ISL_686221\|2020-09 | .................... | .................... | ..................... | .................... | .................... | .................... |
| V | 9224 | hCoV-19/Japan/PG-8626/2020\|EPI_ISL_686222\|2020-09 | .................... | .................... | ..................... | .................... | .................... | .................... |
| V | 9225 | hCoV-19/Japan/PG-8627/2020\|EPI_ISL_686223\|2020-09 | .................... | .................... | ..................... | .................... | .................... | .................... |
| V | 9226 | hCoV-19/Japan/PG-8628/2020\|EPI_ISL_686224\|2020-08 | .................... | .................... | ..................... | .................... | .................... | .................... |
| V | 9227 | hCoV-19/Japan/PG-8629/2020\|EPI_ISL_686225\|2020-08 | .................... | .................... | ..................... | .................... | .................... | .................... |
| V | 9228 | hCoV-19/Japan/PG-8630/2020\|EPI_ISL_686226\|2020-08 | .................... | .................... | ..................... | .................... | .................... | .................... |
| V | 9229 | hCoV-19/Japan/PG-8631/2020\|EPI_ISL_686227\|2020-08 | .................... | .................... | ..................... | .................... | .................... | .................... |
| V | 9230 | hCoV-19/Japan/PG-8632/2020\|EPI_ISL_686228\|2020-08 | .................... | .................... | ..................... | .................... | .................... | .................... |
| V | 9231 | hCoV-19/Japan/PG-8633/2020\|EPI_ISL_686229\|2020-09 | .................... | .................... | ..................... | .................... | .................... | .................... |
| V | 9232 | hCoV-19/Japan/PG-8634/2020\|EPI_ISL_686230\|2020-09 | .................... | .................... | ..................... | .................... | .................... | .................... |
| V | 9233 | hCoV-19/Japan/PG-8635/2020\|EPI_ISL_686231\|2020-09 | .................... | .................... | ..................... | .................... | .................... | .................... |
| V | 9234 | hCoV-19/Japan/PG-8636/2020\|EPI_ISL_686232\|2020-09 | .................... | .................... | ..................... | .................... | .................... | .................... |
| V | 9235 | hCoV-19/Japan/PG-8637/2020\|EPI_ISL_686233\|2020-09 | .................... | .................... | ..................... | .................... | .................... | .................... |
| V | 9236 | hCoV-19/Japan/PG-8638/2020\|EPI_ISL_686234\|2020-09 | .................... | .................... | ..................... | .................... | .................... | .................... |
| V | 9237 | hCoV-19/Japan/PG-8639/2020\|EPI_ISL_686235\|2020-09 | .................... | .................... | ..................... | .................... | .................... | .................... |
| V | 9238 | hCoV-19/Japan/PG-8640/2020\|EPI_ISL_686236\|2020-09 | .................... | .................... | ..................... | .................... | .................... | .................... |
| V | 9239 | hCoV-19/Japan/PG-8641/2020\|EPI_ISL_686237\|2020-09 | .................... | .................... | ..................... | .................... | .................... | .................... |
| V | 9240 | hCoV-19/Japan/PG-8642/2020\|EPI_ISL_686238\|2020-09 | .................... | .................... | ..................... | .................... | .................... | .................... |
| V | 9241 | hCoV-19/Japan/PG-8643/2020\|EPI_ISL_686239\|2020-09 | .................... | .................... | ..................... | .................... | .................... | .................... |
| V | 9242 | hCoV-19/Japan/PG-8644/2020\|EPI_ISL_686240\|2020-09 | .................... | .................... | ..................... | .................... | .................... | .................... |
| V | 9243 | hCoV-19/Japan/PG-8645/2020\|EPI_ISL_686241\|2020-09 | .................... | .................... | ..................... | .................... | .................... | .................... |
| V | 9244 | hCoV-19/Japan/PG-8646/2020\|EPI_ISL_686242\|2020-09 | .................... | .................... | ..................... | .................... | .................... | .................... |
| V | 9245 | hCoV-19/Japan/PG-8647/2020\|EPI_ISL_686243\|2020-09 | .................... | .................... | ..................... | .................... | .................... | .................... |
| V | 9246 | hCoV-19/Japan/PG-8648/2020\|EPI_ISL_686244\|2020-09 | .................... | .................... | ..................... | .................... | .................... | .................... |
| V | 9247 | hCoV-19/Japan/PG-8649/2020\|EPI_ISL_686245\|2020-09 | .................... | .................... | ..................... | .................... | .................... | .................... |
| V | 9248 | hCoV-19/Japan/PG-8650/2020\|EPI_ISL_686246\|2020-09 | .................... | .................... | ..................... | .................... | .................... | .................... |
| V | 9249 | hCoV-19/Japan/PG-8651/2020\|EPI_ISL_686247\|2020-09 | .................... | .................... | ..................... | .................... | .................... | .................... |
| V | 9250 | hCoV-19/Japan/PG-8652/2020\|EPI_ISL_686248\|2020-09 | .................... | .................... | ..................... | .................... | .................... | .................... |
| V | 9251 | hCoV-19/Japan/PG-8653/2020\|EPI_ISL_686249\|2020-09 | .................... | .................... | ..................... | .................... | .................... | .................... |
| V | 9252 | hCoV-19/Japan/PG-8654/2020\|EPI_ISL_686250\|2020-09 | .................... | .................... | ..................... | .................... | .................... | .................... |
| V | 9253 | hCoV-19/Japan/PG-8655/2020\|EPI_ISL_686251\|2020-09 | .................... | .................... | ..................... | .................... | .................... | .................... |
| V | 9254 | hCoV-19/Japan/PG-8656/2020\|EPI_ISL_686252\|2020-09 | .................... | .................... | ..................... | .................... | .................... | .................... |
| V | 9255 | hCoV-19/Japan/PG-8657/2020\|EPI_ISL_686253\|2020-09 | .................... | .................... | ..................... | .................... | .................... | .................... |
| V | 9256 | hCoV-19/Japan/PG-8658/2020\|EPI_ISL_686254\|2020-09 | .................... | .................... | ..................... | .................... | .................... | .................... |
| V | 9257 | hCoV-19/Japan/PG-8659/2020\|EPI_ISL_686255\|2020-09 | .................... | .................... | ..................... | .................... | .................... | .................... |
| V | 9258 | hCoV-19/Japan/PG-8660/2020\|EPI_ISL_686256\|2020-09 | .................... | .................... | ..................... | .................... | .................... | .................... |
| V | 9259 | hCoV-19/Japan/PG-8661/2020\|EPI_ISL_686257\|2020-09 | .................... | .................... | ..................... | .................... | .................... | .................... |
| V | 9260 | hCoV-19/Japan/PG-8662/2020\|EPI_ISL_686258\|2020-09 | .................... | .................... | ..................... | .................... | .................... | .................... |
| V | 9261 | hCoV-19/Japan/PG-8663/2020\|EPI_ISL_686259\|2020-09 | .................... | .................... | ..................... | .................... | .................... | .................... |
| V | 9262 | hCoV-19/Japan/PG-8664/2020\|EPI_ISL_686260\|2020-09 | .................... | .................... | ..................... | .................... | .................... | .................... |
| V | 9263 | hCoV-19/Japan/PG-8665/2020\|EPI_ISL_686261\|2020-09 | .................... | .................... | ..................... | .................... | .................... | .................... |
| V | 9264 | hCoV-19/Japan/PG-8666/2020\|EPI_ISL_686262\|2020-09 | .................... | .................... | ..................... | .................... | .................... | .................... |
| V | 9265 | hCoV-19/Japan/PG-8667/2020\|EPI_ISL_686263\|2020-09 | .................... | .................... | ..................... | .................... | .................... | .................... |
| V | 9266 | hCoV-19/Japan/PG-8668/2020\|EPI_ISL_686264\|2020-09 | .................... | .................... | ..................... | .................... | .................... | .................... |
| V | 9267 | hCoV-19/Japan/PG-8669/2020\|EPI_ISL_686265\|2020-09 | .................... | .................... | ..................... | .................... | .................... | .................... |
| V | 9268 | hCoV-19/Japan/PG-8671/2020\|EPI_ISL_686267\|2020-09 | .................... | .................... | ..................... | .................... | .................... | .................... |
| V | 9269 | hCoV-19/Japan/PG-8672/2020\|EPI_ISL_686268\|2020-09 | .................... | .................... | ..................... | .................... | .................... | .................... |
| V | 9270 | hCoV-19/Japan/PG-8673/2020\|EPI_ISL_686269\|2020-09 | .................... | .................... | ..................... | .................... | .................... | .................... |
| V | 9271 | hCoV-19/Japan/PG-8674/2020\|EPI_ISL_686270\|2020-09 | .................... | .................... | ..................... | .................... | .................... | .................... |
| V | 9272 | hCoV-19/Japan/PG-8675/2020\|EPI_ISL_686271\|2020-09 | .................... | .................... | ..................... | .................... | .................... | .................... |
| V | 9273 | hCoV-19/Japan/PG-8676/2020\|EPI_ISL_686272\|2020-09 | .................... | .................... | ..................... | .................... | .................... | .................... |
| V | 9274 | hCoV-19/Japan/PG-8677/2020\|EPI_ISL_686273\|2020-09 | .................... | .................... | ..................... | .................... | .................... | .................... |
| V | 9275 | hCoV-19/Japan/PG-8678/2020\|EPI_ISL_686274\|2020-09 | .................... | .................... | ..................... | .................... | .................... | .................... |
| V | 9276 | hCoV-19/Japan/PG-8679/2020\|EPI_ISL_686275\|2020-09 | .................... | .................... | ..................... | .................... | .................... | .................... |
| V | 9277 | hCoV-19/Japan/PG-8680/2020\|EPI_ISL_686276\|2020-09 | .................... | .................... | ..................... | .................... | .................... | .................... |
| V | 9278 | hCoV-19/Japan/PG-8681/2020\|EPI_ISL_686277\|2020-09 | .................... | .................... | ..................... | .................... | .................... | .................... |
| V | 9279 | hCoV-19/Japan/PG-8682/2020\|EPI_ISL_686278\|2020-09 | .................... | .................... | ..................... | .................... | .................... | .................... |
| V | 9280 | hCoV-19/Japan/PG-8683/2020\|EPI_ISL_686279\|2020-09 | .................... | .................... | ..................... | .................... | .................... | .................... |
| V | 9281 | hCoV-19/Japan/PG-8684/2020\|EPI_ISL_686280\|2020-09 | .................... | .................... | ..................... | .................... | .................... | .................... |
| V | 9282 | hCoV-19/Japan/PG-8685/2020\|EPI_ISL_686281\|2020-09 | .................... | .................... | ..................... | .................... | .................... | .................... |
| V | 9283 | hCoV-19/Japan/PG-8686/2020\|EPI_ISL_686282\|2020-09 | .................... | .................... | ..................... | .................... | .................... | .................... |
| V | 9284 | hCoV-19/Japan/PG-8687/2020\|EPI_ISL_686283\|2020-08 | .................... | .................... | ..................... | .................... | .................... | .................... |
| V | 9285 | hCoV-19/Japan/PG-8688/2020\|EPI_ISL_686284\|2020-08 | .................... | .................... | ..................... | .................... | .................... | .................... |
| V | 9286 | hCoV-19/Japan/PG-8689/2020\|EPI_ISL_686285\|2020-08 | .................... | .................... | ..................... | .................... | .................... | .................... |
| V | 9287 | hCoV-19/Japan/PG-8690/2020\|EPI_ISL_686286\|2020-08 | .................... | .................... | ..................... | .................... | .................... | .................... |
| V | 9288 | hCoV-19/Japan/PG-8691/2020\|EPI_ISL_686287\|2020-08 | .................... | .................... | ..................... | .................... | .................... | .................... |
| V | 9289 | hCoV-19/Japan/PG-8692/2020\|EPI_ISL_686288\|2020-08 | .................... | .................... | ..................... | .................... | .................... | .................... |
| V | 9290 | hCoV-19/Japan/PG-8693/2020\|EPI_ISL_686289\|2020-08 | .................... | .................... | ..................... | .................... | .................... | .................... |

| | | | | | | | |
|---|---|---|---|---|---|---|---|
| V | 9291 hCoV-19/Japan/PG-8694/2020\|EPI_ISL_686290\|2020-08 | ···················· | ···················· | ····················· | ···················· | ···················· | ···················· |
| V | 9292 hCoV-19/Japan/PG-8695/2020\|EPI_ISL_686291\|2020-08 | ···················· | ···················· | ····················· | ···················· | ···················· | ···················· |
| V | 9293 hCoV-19/Japan/PG-8696/2020\|EPI_ISL_686292\|2020-08 | ···················· | ···················· | ····················· | ···················· | ···················· | ···················· |
| V | 9294 hCoV-19/Japan/PG-8697/2020\|EPI_ISL_686293\|2020-08 | ···················· | ···················· | ····················· | ···················· | ···················· | ···················· |
| V | 9295 hCoV-19/Japan/PG-8698/2020\|EPI_ISL_686294\|2020-08 | ···················· | ···················· | ····················· | ···················· | ···················· | ···················· |
| V | 9296 hCoV-19/Japan/PG-8699/2020\|EPI_ISL_686295\|2020-08 | ···················· | ···················· | ····················· | ···················· | ···················· | ···················· |
| V | 9297 hCoV-19/Japan/PG-8700/2020\|EPI_ISL_686296\|2020-08 | ···················· | ···················· | ····················· | ···················· | ···················· | ···················· |
| V | 9298 hCoV-19/Japan/PG-8701/2020\|EPI_ISL_686297\|2020-08 | ···················· | ···················· | ····················· | ···················· | ···················· | ···················· |
| V | 9299 hCoV-19/Japan/PG-8702/2020\|EPI_ISL_686298\|2020-08 | ···················· | ···················· | ····················· | ···················· | ···················· | ···················· |
| V | 9300 hCoV-19/Japan/PG-8703/2020\|EPI_ISL_686299\|2020-08 | ···················· | ···················· | ····················· | ···················· | ···················· | ···················· |
| V | 9301 hCoV-19/Japan/PG-8447/2020\|EPI_ISL_687546\|2020-09 | ···················· | ···················· | ····················· | ···················· | ···················· | ···················· |
| V | 9302 hCoV-19/Japan/PG-8448/2020\|EPI_ISL_687547\|2020-09 | ···················· | ···················· | ····················· | ···················· | ···················· | ···················· |
| V | 9303 hCoV-19/Japan/PG-8449/2020\|EPI_ISL_687548\|2020-09 | ···················· | ···················· | ····················· | ···················· | ···················· | ···················· |
| V | 9304 hCoV-19/Japan/PG-8450/2020\|EPI_ISL_687549\|2020-09 | ···················· | ··········A········· | ····················· | ···················· | ···················· | ···················· |
| V | 9305 hCoV-19/Japan/PG-8451/2020\|EPI_ISL_687550\|2020-09 | ···················· | ···················· | ····················· | ···················· | ···················· | ···················· |
| V | 9306 hCoV-19/Japan/PG-8452/2020\|EPI_ISL_687551\|2020-09 | ···················· | ···················· | ····················· | ···················· | ···················· | ···················· |
| V | 9307 hCoV-19/Japan/PG-8453/2020\|EPI_ISL_687552\|2020-09 | ···················· | ···················· | ····················· | ···················· | ···················· | ···················· |
| V | 9308 hCoV-19/Japan/PG-8454/2020\|EPI_ISL_687553\|2020-09 | ···················· | ···················· | ····················· | ···················· | ···················· | ···················· |
| V | 9309 hCoV-19/Japan/PG-8455/2020\|EPI_ISL_687554\|2020-09 | ···················· | ···················· | ····················· | ···················· | ···················· | ···················· |
| V | 9310 hCoV-19/Japan/PG-8456/2020\|EPI_ISL_687555\|2020-09 | ···················· | ···················· | ····················· | ···················· | ···················· | ···················· |
| V | 9311 hCoV-19/Japan/PG-8457/2020\|EPI_ISL_687556\|2020-09 | ···················· | ···················· | ····················· | ···················· | ···················· | ···················· |
| V | 9312 hCoV-19/Japan/PG-8458/2020\|EPI_ISL_687557\|2020-09 | ···················· | ···················· | ····················· | ···················· | ···················· | ···················· |
| V | 9313 hCoV-19/Japan/PG-8459/2020\|EPI_ISL_687558\|2020-09 | ···················· | ···················· | ····················· | ···················· | ···················· | ···················· |
| V | 9314 hCoV-19/Japan/PG-8460/2020\|EPI_ISL_687559\|2020-09 | ···················· | ···················· | ···T················· | ···················· | ···················· | ···················· |
| V | 9315 hCoV-19/Japan/PG-8461/2020\|EPI_ISL_687560\|2020-09 | ···················· | ···················· | ····················· | ···················· | ···················· | ···················· |
| V | 9316 hCoV-19/Japan/PG-8462/2020\|EPI_ISL_687561\|2020-06 | ···················· | ···················· | ····················· | ···················· | ···················· | ···················· |
| V | 9317 hCoV-19/Japan/PG-8463/2020\|EPI_ISL_687562\|2020-07 | ···················· | ···················· | ····················· | ···················· | ···················· | ···················· |
| V | 9318 hCoV-19/Japan/PG-8464/2020\|EPI_ISL_687563\|2020-07 | ···················· | ···················· | ····················· | ···················· | ···················· | ···················· |
| V | 9319 hCoV-19/Japan/PG-8465/2020\|EPI_ISL_687564\|2020-08 | ···················· | ···················· | ····················· | ···················· | ···················· | ···················· |
| V | 9320 hCoV-19/Japan/PG-8466/2020\|EPI_ISL_687565\|2020-08 | ···················· | ···················· | ····················· | ···················· | ···················· | ···················· |
| V | 9321 hCoV-19/Japan/PG-8467/2020\|EPI_ISL_687566\|2020-08 | ···················· | ···················· | ····················· | ···················· | ···················· | ···················· |
| V | 9322 hCoV-19/Japan/PG-8468/2020\|EPI_ISL_687567\|2020-06 | ···················· | ···················· | ····················· | ···················· | ···················· | ···················· |
| V | 9323 hCoV-19/Japan/PG-8469/2020\|EPI_ISL_687568\|2020-06 | ···················· | ···················· | ····················· | ···················· | ···················· | ···················· |
| V | 9324 hCoV-19/Japan/PG-8470/2020\|EPI_ISL_687569\|2020-07 | ···················· | ···················· | ····················· | ···················· | ···················· | ···················· |
| V | 9325 hCoV-19/Japan/PG-8471/2020\|EPI_ISL_687570\|2020-08 | ···················· | ···················· | ····················· | ···················· | ···················· | ···················· |
| V | 9326 hCoV-19/Japan/PG-8473/2020\|EPI_ISL_687572\|2020-07 | ···················· | ···················· | ····················· | ···················· | ···················· | ···················· |
| V | 9327 hCoV-19/Japan/PG-8474/2020\|EPI_ISL_687573\|2020-08 | ···················· | ···················· | ····················· | ···················· | ···················· | ···················· |
| V | 9328 hCoV-19/Japan/PG-8475/2020\|EPI_ISL_687574\|2020-08 | ···················· | ···················· | ····················· | ···················· | ···················· | ···················· |
| V | 9329 hCoV-19/Japan/PG-8476/2020\|EPI_ISL_687575\|2020-08 | ···················· | ···················· | ····················· | ···················· | ···················· | ···················· |
| V | 9330 hCoV-19/Japan/PG-8477/2020\|EPI_ISL_687576\|2020-08 | ···················· | ···················· | ····················· | ···················· | ···················· | ···················· |
| V | 9331 hCoV-19/Japan/PG-8478/2020\|EPI_ISL_687577\|2020-07 | ···················· | ···················· | ····················· | ···················· | ···················· | ···················· |
| V | 9332 hCoV-19/Japan/PG-8479/2020\|EPI_ISL_687578\|2020-08 | ···················· | ···················· | ····················· | ···················· | ···················· | ···················· |
| V | 9333 hCoV-19/Japan/PG-8480/2020\|EPI_ISL_687579\|2020-08 | ···················· | ···················· | ····················· | ···················· | ···················· | ···················· |
| V | 9334 hCoV-19/Japan/PG-8481/2020\|EPI_ISL_687580\|2020-08 | ···················· | ···················· | ····················· | ···················· | ···················· | ···················· |
| V | 9335 hCoV-19/Japan/PG-8482/2020\|EPI_ISL_687581\|2020-08 | ···················· | ···················· | ····················· | ···················· | ···················· | ···················· |
| V | 9336 hCoV-19/Japan/PG-8483/2020\|EPI_ISL_687582\|2020-08 | ···················· | ···················· | ····················· | ···················· | ···················· | ···················· |
| V | 9337 hCoV-19/Japan/PG-8484/2020\|EPI_ISL_687583\|2020-08 | ···················· | ···················· | ····················· | ···················· | ···················· | ···················· |
| V | 9338 hCoV-19/Japan/PG-8485/2020\|EPI_ISL_687584\|2020-06 | ···················· | ···················· | ····················· | ···················· | ···················· | ···················· |
| V | 9339 hCoV-19/Japan/PG-8486/2020\|EPI_ISL_687585\|2020-06 | ···················· | ···················· | ····················· | ···················· | ···················· | ···················· |
| V | 9340 hCoV-19/Japan/PG-8487/2020\|EPI_ISL_687586\|2020-06 | ···················· | ···················· | ····················· | ···················· | ···················· | ···················· |
| V | 9341 hCoV-19/Japan/PG-8488/2020\|EPI_ISL_687587\|2020-06 | ···················· | ···················· | ····················· | ···················· | ···················· | ···················· |
| V | 9342 hCoV-19/Japan/PG-8489/2020\|EPI_ISL_687588\|2020-06 | ···················· | ···················· | ····················· | ···················· | ···················· | ···················· |
| V | 9343 hCoV-19/Japan/PG-8490/2020\|EPI_ISL_687589\|2020-06 | ···················· | ···················· | ····················· | ···················· | ···················· | ···················· |
| V | 9344 hCoV-19/Japan/PG-8491/2020\|EPI_ISL_687590\|2020-06 | ···················· | ···················· | ····················· | ···················· | ···················· | ···················· |
| V | 9345 hCoV-19/Japan/PG-8492/2020\|EPI_ISL_687591\|2020-06 | ···················· | ···················· | ····················· | ···················· | ···················· | ···················· |
| V | 9346 hCoV-19/Japan/PG-8493/2020\|EPI_ISL_687592\|2020-06 | ···················· | ···················· | ····················· | ···················· | ···················· | ···················· |
| V | 9347 hCoV-19/Japan/PG-8494/2020\|EPI_ISL_687593\|2020-06 | ···················· | ···················· | ····················· | ···················· | ···················· | ···················· |
| V | 9348 hCoV-19/Japan/PG-8495/2020\|EPI_ISL_687594\|2020-06 | ···················· | ···················· | ····················· | ···················· | ···················· | ···················· |
| V | 9349 hCoV-19/Japan/PG-8496/2020\|EPI_ISL_687595\|2020-06 | ···················· | ···················· | ····················· | ···················· | ···················· | ···················· |
| V | 9350 hCoV-19/Japan/PG-8497/2020\|EPI_ISL_687596\|2020-06 | ···················· | ···················· | ····················· | ···················· | ···················· | ···················· |
| V | 9351 hCoV-19/Japan/PG-8498/2020\|EPI_ISL_687597\|2020-06 | ···················· | ···················· | ····················· | ···················· | ···················· | ···················· |
| V | 9352 hCoV-19/Japan/PG-8499/2020\|EPI_ISL_687598\|2020-06 | ···················· | ···················· | ····················· | ···················· | ···················· | ···················· |
| V | 9353 hCoV-19/Japan/PG-8500/2020\|EPI_ISL_687599\|2020-07 | ···················· | ···················· | ····················· | ···················· | ···················· | ···················· |
| V | 9354 hCoV-19/Japan/PG-5758/2020\|EPI_ISL_688877\|2020-08 | ···················· | ···················· | ····················· | ···················· | ···················· | ···················· |
| V | 9355 hCoV-19/Japan/PG-5759/2020\|EPI_ISL_688878\|2020-08 | ···················· | ···················· | ····················· | ···················· | ···················· | ···················· |
| V | 9356 hCoV-19/Japan/PG-5760/2020\|EPI_ISL_688879\|2020-08 | ···················· | ···················· | ····················· | ···················· | ···················· | ···················· |
| V | 9357 hCoV-19/Japan/PG-5761/2020\|EPI_ISL_688880\|2020-08 | ···················· | ···················· | ····················· | ···················· | ···················· | ···················· |
| V | 9358 hCoV-19/Japan/PG-5762/2020\|EPI_ISL_688881\|2020-08 | ···················· | ···················· | ····················· | ···················· | ···················· | ···················· |
| V | 9359 hCoV-19/Japan/PG-5763/2020\|EPI_ISL_688882\|2020-08 | ···················· | ···················· | ····················· | ···················· | ···················· | ···················· |
| V | 9360 hCoV-19/Japan/PG-5764/2020\|EPI_ISL_688883\|2020-08 | ···················· | ···················· | ····················· | ···················· | ···················· | ···················· |
| V | 9361 hCoV-19/Japan/PG-5765/2020\|EPI_ISL_688884\|2020-08 | ···················· | ···················· | ····················· | ···················· | ···················· | ···················· |
| V | 9362 hCoV-19/Japan/PG-5766/2020\|EPI_ISL_688885\|2020-08 | ···················· | ···················· | ····················· | ···················· | ···················· | ···················· |
| V | 9363 hCoV-19/Japan/PG-5767/2020\|EPI_ISL_688886\|2020-08 | ···················· | ···················· | ····················· | ···················· | ···················· | ···················· |
| V | 9364 hCoV-19/Japan/PG-5768/2020\|EPI_ISL_688887\|2020-07 | ···················· | ···················· | ····················· | ···················· | ···················· | ···················· |
| V | 9365 hCoV-19/Japan/PG-5769/2020\|EPI_ISL_688888\|2020-07 | ···················· | ···················· | ····················· | ···················· | ···················· | ···················· |
| V | 9366 hCoV-19/Japan/PG-5770/2020\|EPI_ISL_688889\|2020-07 | ···················· | ···················· | ····················· | ···················· | ···················· | ···················· |
| V | 9367 hCoV-19/Japan/PG-5771/2020\|EPI_ISL_688890\|2020-07 | ···················· | ···················· | ····················· | ···················· | ···················· | ···················· |
| V | 9368 hCoV-19/Japan/PG-5772/2020\|EPI_ISL_688891\|2020-07 | ···················· | ···················· | ····················· | ···················· | ···················· | ···················· |
| V | 9369 hCoV-19/Japan/PG-5773/2020\|EPI_ISL_688892\|2020-07 | ···················· | ···················· | ····················· | ···················· | ···················· | ···················· |
| V | 9370 hCoV-19/Japan/PG-5774/2020\|EPI_ISL_688893\|2020-07 | ···················· | ···················· | ····················· | ···················· | ···················· | ···················· |

| | | | | | | | | |
|---|---|---|---|---|---|---|---|---|
| V | 9371 | hCoV-19/Japan/PG-5775/2020\|EPI_ISL_688894\|2020-08 | ···················· | ···················· | ····················· | ···················· | ···················· | ···················· |
| V | 9372 | hCoV-19/Japan/PG-5776/2020\|EPI_ISL_688895\|2020-08 | ···················· | ···················· | ····················· | ···················· | ···················· | ···················· |
| V | 9373 | hCoV-19/Japan/PG-5777/2020\|EPI_ISL_688896\|2020-08 | ···················· | ···················· | ····················· | ···················· | ···················· | ···················· |
| V | 9374 | hCoV-19/Japan/PG-5778/2020\|EPI_ISL_688897\|2020-08 | ···················· | ···················· | ····················· | ···················· | ···················· | ···················· |
| V | 9375 | hCoV-19/Japan/PG-5779/2020\|EPI_ISL_688898\|2020-08 | ···················· | ···················· | ····················· | ···················· | ···················· | ···················· |
| V | 9376 | hCoV-19/Japan/PG-5780/2020\|EPI_ISL_688899\|2020-08 | ···················· | ···················· | ····················· | ···················· | ···················· | ···················· |
| V | 9377 | hCoV-19/Japan/PG-5791/2020\|EPI_ISL_688910\|2020-08 | ···················· | ···················· | ····················· | ···················· | ···················· | ···················· |
| V | 9378 | hCoV-19/Japan/PG-5792/2020\|EPI_ISL_688911\|2020-08 | ···················· | ···················· | ····················· | ···················· | ···················· | ···················· |
| V | 9379 | hCoV-19/Japan/PG-5793/2020\|EPI_ISL_688912\|2020-08 | ···················· | ···················· | ····················· | ···················· | ···················· | ···················· |
| V | 9380 | hCoV-19/Japan/PG-5794/2020\|EPI_ISL_688913\|2020-08 | ···················· | ···················· | ····················· | ···················· | ···················· | ···················· |
| V | 9381 | hCoV-19/Japan/PG-5795/2020\|EPI_ISL_688914\|2020-08 | ···················· | ···················· | ····················· | ···················· | ···················· | ···················· |
| V | 9382 | hCoV-19/Japan/PG-5796/2020\|EPI_ISL_688915\|2020-08 | ···················· | ···················· | ····················· | ···················· | ···················· | ···················· |
| V | 9383 | hCoV-19/Japan/PG-5797/2020\|EPI_ISL_688916\|2020-08 | ···················· | ···················· | ····················· | ···················· | ···················· | ···················· |
| V | 9384 | hCoV-19/Japan/PG-5798/2020\|EPI_ISL_688917\|2020-08 | ···················· | ···················· | ····················· | ···················· | ···················· | ···················· |
| V | 9385 | hCoV-19/Japan/PG-5799/2020\|EPI_ISL_688918\|2020-08 | ···················· | ···················· | ····················· | ···················· | ···················· | ···················· |
| V | 9386 | hCoV-19/Japan/PG-5800/2020\|EPI_ISL_688919\|2020-08 | ···················· | ···················· | ····················· | ···················· | ···················· | ···················· |
| V | 9387 | hCoV-19/Japan/PG-5802/2020\|EPI_ISL_688920\|2020-08 | ···················· | ···················· | ····················· | ···················· | ···················· | ···················· |
| V | 9388 | hCoV-19/Japan/PG-5803/2020\|EPI_ISL_688921\|2020-08 | ···················· | ···················· | ····················· | ···················· | ···················· | ···················· |
| V | 9389 | hCoV-19/Japan/PG-5804/2020\|EPI_ISL_688922\|2020-08 | ···················· | ···················· | ····················· | ···················· | ···················· | ···················· |
| V | 9390 | hCoV-19/Japan/PG-5805/2020\|EPI_ISL_688923\|2020-08 | ···················· | ···················· | ····················· | ···················· | ···················· | ···················· |
| V | 9391 | hCoV-19/Japan/PG-5806/2020\|EPI_ISL_688924\|2020-08 | ···················· | ···················· | ····················· | ···················· | ···················· | ···················· |
| V | 9392 | hCoV-19/Japan/PG-5807/2020\|EPI_ISL_688925\|2020-08 | ···················· | ···················· | ····················· | ···················· | ···················· | ···················· |
| V | 9393 | hCoV-19/Japan/PG-5809/2020\|EPI_ISL_688926\|2020-08 | ···················· | ···················· | ····················· | ···················· | ···················· | ···················· |
| V | 9394 | hCoV-19/Japan/PG-5810/2020\|EPI_ISL_688927\|2020-08 | ···················· | ···················· | ····················· | ···················· | ···················· | ···················· |
| V | 9395 | hCoV-19/Japan/PG-5811/2020\|EPI_ISL_688928\|2020-08 | ···················· | ···················· | ····················· | ···················· | ···················· | ···················· |
| V | 9396 | hCoV-19/Japan/PG-3915/2020\|EPI_ISL_692166\|2020-07 | ···················· | ···················· | ····················· | ···················· | ···················· | ···················· |
| V | 9397 | hCoV-19/Japan/PG-3916/2020\|EPI_ISL_692167\|2020-07 | ···················· | ···················· | ····················· | ···················· | ···················· | ···················· |
| V | 9398 | hCoV-19/Japan/PG-3917/2020\|EPI_ISL_692168\|2020-07 | ···················· | ···················· | ····················· | ···················· | ···················· | ···················· |
| V | 9399 | hCoV-19/Japan/PG-3918/2020\|EPI_ISL_692169\|2020-07 | ···················· | ···················· | ····················· | ···················· | ···················· | ···················· |
| V | 9400 | hCoV-19/Japan/PG-3919/2020\|EPI_ISL_692170\|2020-07 | ···················· | ···················· | ····················· | ···················· | ···················· | ···················· |
| V | 9401 | hCoV-19/Japan/PG-3920/2020\|EPI_ISL_692171\|2020-07 | ···················· | ···················· | ····················· | ···················· | ···················· | ···················· |
| V | 9402 | hCoV-19/Japan/PG-3921/2020\|EPI_ISL_692172\|2020-07 | ···················· | ···················· | ····················· | ···················· | ···················· | ···················· |
| V | 9403 | hCoV-19/Japan/PG-3922/2020\|EPI_ISL_692173\|2020-07 | ···················· | ···················· | ····················· | ···················· | ···················· | ···················· |
| V | 9404 | hCoV-19/Japan/PG-3923/2020\|EPI_ISL_692174\|2020-07 | ···················· | ···················· | ····················· | ···················· | ···················· | ···················· |
| V | 9405 | hCoV-19/Japan/PG-3924/2020\|EPI_ISL_692175\|2020-07 | ···················· | ···················· | ····················· | ···················· | ···················· | ···················· |
| V | 9406 | hCoV-19/Japan/PG-3925/2020\|EPI_ISL_692176\|2020-07 | ···················· | ···················· | ····················· | ···················· | ···················· | ···················· |
| V | 9407 | hCoV-19/England/204740534/2020\|EPI_ISL_693406\|2020 | ···················· | ···················· | ····················· | ···················· | ···················· | ···················· |
| V | 9408 | hCoV-19/England/204740524/2020\|EPI_ISL_693407\|2020 | ···················· | ···················· | ····················· | ···················· | ···················· | ···················· |
| V | 9409 | hCoV-19/England/204661316/2020\|EPI_ISL_693408\|2020 | ···················· | ···················· | ····················· | ···················· | ···················· | ···················· |
| V | 9410 | hCoV-19/England/204661328/2020\|EPI_ISL_693409\|2020 | ···················· | ···················· | ····················· | ···················· | ···················· | ···················· |
| V | 9411 | hCoV-19/England/204661353/2020\|EPI_ISL_693410\|2020 | ···················· | ···················· | ····················· | ···················· | ···················· | ···················· |
| V | 9412 | hCoV-19/England/204661424/2020\|EPI_ISL_693411\|2020 | ···················· | ···················· | ····················· | ···················· | ···················· | ···················· |
| V | 9413 | hCoV-19/England/204661580/2020\|EPI_ISL_693412\|2020 | ···················· | ···················· | ····················· | ···················· | ···················· | ···················· |
| V | 9414 | hCoV-19/England/204661581/2020\|EPI_ISL_693413\|2020 | ···················· | ···················· | ····················· | ···················· | ······T············· | ···················· |
| V | 9415 | hCoV-19/England/204661582/2020\|EPI_ISL_693414\|2020 | ···················· | ···················· | ····················· | ···················· | ······T············· | ···················· |
| V | 9416 | hCoV-19/England/204661601/2020\|EPI_ISL_693415\|2020 | ···················· | ···················· | ····················· | ···················· | ···················· | ···················· |
| V | 9417 | hCoV-19/England/204661604/2020\|EPI_ISL_693416\|2020 | ···················· | ···················· | ····················· | ···················· | ···················· | ···················· |
| V | 9418 | hCoV-19/England/204661640/2020\|EPI_ISL_693417\|2020 | ···················· | ···················· | ····················· | ···················· | ······T············· | ···················· |
| V | 9419 | hCoV-19/England/204682055/2020\|EPI_ISL_693418\|2020 | ···················· | ···················· | ····················· | ···················· | ···················· | ···················· |
| V | 9420 | hCoV-19/England/204682063/2020\|EPI_ISL_693419\|2020 | ···················· | ···················· | ····················· | ···················· | ···················· | ···················· |
| V | 9421 | hCoV-19/France/ARA-72179/2020\|EPI_ISL_693497\|2020- | ···················· | ···················· | ····················· | ···················· | ···················· | ···················· |
| V | 9422 | hCoV-19/France/ARA-72266/2020\|EPI_ISL_693498\|2020- | ···················· | ···················· | ····················· | ···················· | ···················· | ···················· |
| V | 9423 | hCoV-19/France/ARA-72824/2020\|EPI_ISL_693499\|2020- | ···················· | ···················· | ····················· | ···················· | ···················· | ···················· |
| W | 1 | hCoV-19/USA/AZ-TG489927/2020\|EPI_ISL_695100\|2020-0 | ···················· | ···················· | ····················· | ···················· | ···················· | ···················· |
| W | 2 | hCoV-19/USA/AZ-TG491468/2020\|EPI_ISL_695103\|2020-0 | ···················· | ···················· | ····················· | ···················· | ···················· | ···················· |
| W | 3 | hCoV-19/USA/AZ-TG491469/2020\|EPI_ISL_695104\|2020-0 | ···················· | ······A············· | ····················· | ···················· | ···················· | ···················· |
| W | 4 | hCoV-19/USA/AZ-TG491470/2020\|EPI_ISL_695105\|2020-0 | ···················· | ···················· | ····················· | ···················· | ···················· | ···················· |
| W | 5 | hCoV-19/USA/AZ-TG491471/2020\|EPI_ISL_695106\|2020-0 | ···················· | ···················· | ····················· | ···················· | ···················· | ···················· |
| W | 6 | hCoV-19/USA/AZ-TG491475/2020\|EPI_ISL_695109\|2020-0 | ···················· | ···················· | ····················· | ···················· | ···················· | ···················· |
| W | 7 | hCoV-19/USA/AZ-TG491477/2020\|EPI_ISL_695110\|2020-0 | ···················· | ···················· | ····················· | ···················· | ···················· | ···················· |
| W | 8 | hCoV-19/USA/AZ-TG491478/2020\|EPI_ISL_695111\|2020-0 | ···················· | ···················· | ····················· | ···················· | ···················· | ···················· |
| W | 9 | hCoV-19/USA/AZ-TG491479/2020\|EPI_ISL_695112\|2020-0 | ···················· | ·A·················· | ····················· | ···················· | ···················· | ···················· |
| W | 10 | hCoV-19/USA/AZ-TG491480/2020\|EPI_ISL_695113\|2020-0 | ···················· | ···················· | ····················· | ···················· | ···················· | ···················· |
| W | 11 | hCoV-19/USA/AZ-TG491482/2020\|EPI_ISL_695114\|2020-0 | ···················· | ···················· | ····················· | ···················· | ···················· | ···················· |
| W | 12 | hCoV-19/USA/AZ-TG491484/2020\|EPI_ISL_695116\|2020-0 | ···················· | ·A·················· | ····················· | ···················· | ···················· | ···················· |
| W | 13 | hCoV-19/USA/AZ-TG491485/2020\|EPI_ISL_695117\|2020-0 | ···················· | ·A·················· | ····················· | ···················· | ···················· | ···················· |
| W | 14 | hCoV-19/USA/AZ-TG491487/2020\|EPI_ISL_695118\|2020-0 | ···················· | ·A·················· | ····················· | ···················· | ···················· | ···················· |
| W | 15 | hCoV-19/USA/AZ-TG491488/2020\|EPI_ISL_695119\|2020-0 | ···················· | ···················· | ····················· | ···················· | ···················· | ···················· |
| W | 16 | hCoV-19/USA/AZ-TG491490/2020\|EPI_ISL_695121\|2020-0 | ···················· | ···················· | ····················· | ···················· | ···················· | ···················· |
| W | 17 | hCoV-19/USA/AZ-TG491494/2020\|EPI_ISL_695124\|2020-0 | ···················· | ·A·················· | ····················· | ···················· | ···················· | ···················· |
| W | 18 | hCoV-19/USA/AZ-TG491498/2020\|EPI_ISL_695127\|2020-0 | ···················· | ···················· | ····················· | ···················· | ···················· | ···················· |
| W | 19 | hCoV-19/USA/AZ-TG491500/2020\|EPI_ISL_695129\|2020-0 | ···················· | ·N·················· | ····················· | ···················· | ···················· | ···················· |
| W | 20 | hCoV-19/USA/AZ-TG491502/2020\|EPI_ISL_695130\|2020-0 | ···················· | ···················· | ····················· | ···················· | ···················· | ···················· |
| W | 21 | hCoV-19/USA/AZ-TG491507/2020\|EPI_ISL_695131\|2020-0 | ···················· | ···················· | ····················· | ···················· | ···················· | ···················· |
| W | 22 | hCoV-19/USA/AZ-TG491508/2020\|EPI_ISL_695132\|2020-0 | ···················· | ···················· | ····················· | ···················· | ···················· | ···················· |
| W | 23 | hCoV-19/USA/AZ-TG491509/2020\|EPI_ISL_695133\|2020-0 | ···················· | ···················· | ····················· | ···················· | ···················· | ···················· |
| W | 24 | hCoV-19/USA/AZ-TG491510/2020\|EPI_ISL_695134\|2020-0 | ···················· | ·············A······ | ····················· | ···················· | ···················· | ···················· |
| W | 25 | hCoV-19/USA/AZ-TG491511/2020\|EPI_ISL_695135\|2020-0 | ···················· | ·A·················· | ····················· | ···················· | ···················· | ···················· |
| W | 26 | hCoV-19/USA/AZ-TG491512/2020\|EPI_ISL_695136\|2020-0 | ···················· | ·A·················· | ····················· | ···················· | ···················· | ···················· |
| W | 27 | hCoV-19/USA/AZ-TG491513/2020\|EPI_ISL_695137\|2020-0 | ···················· | ·A·················· | ····················· | ···················· | ···················· | ···················· |

```
W    28 hCoV-19/USA/AZ-TG491514/2020|EPI_ISL_695138|2020-0   ····················  ·A··················  ·····················  ····················  ····················  ····················
W    29 hCoV-19/USA/AZ-TG491515/2020|EPI_ISL_695139|2020-0   ····················  ····················  ·····················  ····················  ····················  ····················
W    30 hCoV-19/USA/AZ-TG491526/2020|EPI_ISL_695144|2020-0   ····················  ····················  ·····················  ····················  ····················  ····················
W    31 hCoV-19/USA/AZ-TG491528/2020|EPI_ISL_695145|2020-0   ····················  ····················  ·····················  ····················  ····················  ····················
W    32 hCoV-19/USA/AZ-TG491530/2020|EPI_ISL_695147|2020-0   ····················  ····················  ·····················  ····················  ····················  ····················
W    33 hCoV-19/USA/AZ-TG491531/2020|EPI_ISL_695148|2020-0   ····················  ····················  ·····················  ····················  ····················  ····················
W    34 hCoV-19/USA/AZ-TG491533/2020|EPI_ISL_695150|2020-0   ····················  ····················  ·····················  ····················  ····················  ····················
W    35 hCoV-19/USA/AZ-TG491534/2020|EPI_ISL_695151|2020-0   ····················  ····················  ·····················  ····················  ····················  ····················
W    36 hCoV-19/USA/AZ-TG491535/2020|EPI_ISL_695152|2020-0   ····················  ····················  ·····················  ····················  ····················  ····················
W    37 hCoV-19/USA/AZ-TG491537/2020|EPI_ISL_695154|2020-0   ····················  ····················  ·····················  ····················  ····················  ····················
W    38 hCoV-19/USA/AZ-TG491538/2020|EPI_ISL_695155|2020-0   ····················  ····················  ·····················  ····················  ····················  ····················
W    39 hCoV-19/USA/AZ-TG491542/2020|EPI_ISL_695158|2020-0   ····················  ····················  ·····················  ····················  ····················  ····················
W    40 hCoV-19/USA/AZ-TG491543/2020|EPI_ISL_695159|2020-0   ····················  ····················  ·····················  ····················  ····················  ····················
W    41 hCoV-19/USA/AZ-TG491544/2020|EPI_ISL_695160|2020-0   ····················  ····················  ·····················  ····················  ····················  ····················
W    42 hCoV-19/USA/AZ-TG491547/2020|EPI_ISL_695161|2020-0   ····················  ····················  ·····················  ····················  ····················  ····················
W    43 hCoV-19/USA/AZ-TG491549/2020|EPI_ISL_695163|2020-0   ····················  ····················  ·····················  ····················  ····················  ····················
W    44 hCoV-19/USA/AZ-TG491551/2020|EPI_ISL_695165|2020-0   ····················  ·A··················  ·····················  ····················  ····················  ····················
W    45 hCoV-19/USA/AZ-TG491552/2020|EPI_ISL_695166|2020-0   ····················  ·A··················  ·····················  ····················  ····················  ····················
W    46 hCoV-19/USA/AZ-TG491553/2020|EPI_ISL_695167|2020-0   ····················  ····················  ·····················  ····················  ····················  ····················
W    47 hCoV-19/USA/AZ-TG670230/2020|EPI_ISL_696412|2020-0   ····················  ····················  ·····················  ····················  ····················  ····················
W    48 hCoV-19/USA/AZ-TG670233/2020|EPI_ISL_696414|2020-0   ····················  ····················  ·····················  ····················  ····················  ····················
W    49 hCoV-19/USA/AZ-TG670236/2020|EPI_ISL_696416|2020-0   ····················  ····················  ·····················  ····················  ····················  ····················
W    50 hCoV-19/USA/AZ-TG670240/2020|EPI_ISL_696420|2020-0   ····················  ····················  ·····················  ····················  ····················  ····················
W    51 hCoV-19/USA/AZ-TG670252/2020|EPI_ISL_696430|2020-0   ····················  ····················  ·····················  ····················  ····················  ····················
W    52 hCoV-19/USA/AZ-TG670255/2020|EPI_ISL_696432|2020-0   ····················  ····················  ·····················  ····················  ····················  ····················
W    53 hCoV-19/USA/AZ-TG670257/2020|EPI_ISL_696433|2020-0   ····················  ····················  ·····················  ····················  ····················  ····················
W    54 hCoV-19/USA/AZ-TG670268/2020|EPI_ISL_696443|2020-0   ····················  ····················  ·····················  ····················  ····················  ····················
W    55 hCoV-19/USA/AZ-TG670276/2020|EPI_ISL_696447|2020-0   ····················  ····················  ·····················  ····················  ····················  ····················
W    62 hCoV-19/South Africa/NHLS-UCT-GS-1671-KRISP/2020|E   ····················  ····················  ·····················  ····················  ····················  ····················
W    64 hCoV-19/South Africa/NHLS-UCT-GS-1391-KRISP/2020|E   ····················  ·A··················  ·····················  ····················  ····················  ····················
W    66 hCoV-19/South Africa/NHLS-UCT-GS-7004-KRISP/2020|E   ····················  ·A··················  ·····················  ····················  ····················  ····················
W    83 hCoV-19/Ecuador/USFQ-173/2020|EPI_ISL_697783|2020-   ····················  ····················  ·····················  ····················  ····················  ····················
W    84 hCoV-19/Ecuador/USFQ-185/2020|EPI_ISL_697784|2020-   ····················  ····················  ·····················  ····················  ····················  ····················
W    85 hCoV-19/Ecuador/USFQ-1188/2020|EPI_ISL_697785|2020   ····················  ····················  ·····················  ····················  ····················  ····················
W    86 hCoV-19/Ecuador/USFQ-189/2020|EPI_ISL_697786|2020-   ····················  ····················  ·····················  ····················  ····················  ····················
W    87 hCoV-19/Ecuador/USFQ-322/2020|EPI_ISL_697788|2020-   ····················  ····················  ·····················  ····················  ····················  ····················
W    88 hCoV-19/Ecuador/USFQ-344/2020|EPI_ISL_697789|2020-   ····················  ····················  ·····················  ····················  ····················  ····················
W    89 hCoV-19/Ecuador/USFQ-346/2020|EPI_ISL_697790|2020-   ····················  ····················  ·····················  ····················  ····················  ····················
W    90 hCoV-19/Ecuador/USFQ-405/2020|EPI_ISL_697792|2020-   ····················  ····················  ·····················  ····················  ····················  ····················
W    91 hCoV-19/Ecuador/USFQ-433/2020|EPI_ISL_697793|2020-   ····················  ····················  ·····················  ····················  ····················  ····················
W    92 hCoV-19/Ecuador/USFQ-523/2020|EPI_ISL_697794|2020-   ····················  ····················  ·····················  ····················  ····················  ····················
W    93 hCoV-19/Ecuador/USFQ-524/2020|EPI_ISL_697795|2020-   ····················  ····················  ·····················  ····················  ····················  ····················
W    94 hCoV-19/Ecuador/USFQ-527/2020|EPI_ISL_697796|2020-   ····················  ····················  ·····················  ····················  ····················  ····················
W    95 hCoV-19/Ecuador/USFQ-531/2020|EPI_ISL_697798|2020-   ····················  ····················  ·····················  ····················  ····················  ····················
W    96 hCoV-19/Ecuador/USFQ-533/2020|EPI_ISL_697799|2020-   ····················  ····················  ·····················  ····················  ····················  ····················
W    97 hCoV-19/England/ALDP-BB2A40/2020|EPI_ISL_702800|20   ····················  ····················  ·····················  ····················  ····················  ····················
W    98 hCoV-19/England/ALDP-BB2B4D/2020|EPI_ISL_702802|20   ····················  ····················  ·····················  ····················  ····················  ····················
W    99 hCoV-19/England/CAMC-BC0845/2020|EPI_ISL_705003|20   ····················  ·A··················  ·····················  ····················  ····················  ····················
W   100 hCoV-19/England/CAMC-BC072A/2020|EPI_ISL_705006|20   ····················  ·A··················  ·····················  ····················  ····················  ···A················
W   101 hCoV-19/England/CAMC-BC0739/2020|EPI_ISL_705009|20   ····················  ····················  ·····················  ····················  ····················  ····················
W   102 hCoV-19/England/CAMC-BC1F8B/2020|EPI_ISL_705011|20   ····················  ····················  ·····················  ····················  ····················  ····················
W   103 hCoV-19/England/CAMC-BC0D46/2020|EPI_ISL_705012|20   ····················  ·A··················  ·····················  ····················  ····················  ····················
W   104 hCoV-19/England/CAMC-BC1FA9/2020|EPI_ISL_705014|20   ····················  ····················  ·····················  ····················  ····················  ····················
W   105 hCoV-19/England/CAMC-BC0A6D/2020|EPI_ISL_705016|20   ····················  ····················  ·····················  ····················  ····················  ····················
W   106 hCoV-19/England/CAMC-BC012C/2020|EPI_ISL_705025|20   ····················  ····················  ·····················  ····················  ····················  ····················
W   107 hCoV-19/England/CAMC-BC070C/2020|EPI_ISL_705027|20   ····················  ····················  ·····················  ····················  ······T·············  ····················
W   108 hCoV-19/England/CAMC-BC0B3D/2020|EPI_ISL_705028|20   ····················  ····················  ·····················  ····················  ····T···············  ····················
W   109 hCoV-19/England/CAMC-BC0881/2020|EPI_ISL_705031|20   ····················  ····················  ·····················  ····················  ····················  ····················
W   110 hCoV-19/England/CAMC-BC22EB/2020|EPI_ISL_705034|20   ····················  ····················  ·····················  ····················  ····················  ····················
W   111 hCoV-19/England/CAMC-BC0AF4/2020|EPI_ISL_705036|20   ····················  ····················  ·····················  ····················  ····················  ····················
W   112 hCoV-19/England/CAMC-BC0C3A/2020|EPI_ISL_705037|20   ····················  ····················  ·····················  ····················  ····················  ····················
W   113 hCoV-19/England/CAMC-BC0A8B/2020|EPI_ISL_705041|20   ····················  ····················  ·····················  ····················  ····················  ····················
W   114 hCoV-19/England/CAMC-BC0C85/2020|EPI_ISL_705042|20   ····················  ····················  ·····················  ····················  ····················  ····················
W   115 hCoV-19/England/CAMC-BC0344/2020|EPI_ISL_705045|20   ····················  ····················  ·····················  ····················  ····················  ····················
W   116 hCoV-19/England/CAMC-BC0872/2020|EPI_ISL_705047|20   ····················  ·A··················  ·····················  ····················  ····················  ····················
W   117 hCoV-19/England/CAMC-BC0BE2/2020|EPI_ISL_705049|20   ····················  ····················  ·····················  ····················  ····················  ····················
W   118 hCoV-19/England/CAMC-BC06C3/2020|EPI_ISL_705050|20   ····················  ····················  ·····················  ····················  ····················  ····················
W   119 hCoV-19/England/CAMC-BC0326/2020|EPI_ISL_705052|20   ····················  ····················  ·····················  ····················  ····················  ····················
W   120 hCoV-19/England/CAMC-BBFEED/2020|EPI_ISL_705053|20   ····················  ·A··················  ·····················  ····················  ····················  ····················
W   121 hCoV-19/England/CAMC-BC0A30/2020|EPI_ISL_705060|20   ····················  ····················  ·····················  ····················  ····················  ····················
W   122 hCoV-19/England/CAMC-BC110D/2020|EPI_ISL_705066|20   ····················  ····················  ·····················  ····················  ····················  ····················
W   123 hCoV-19/England/CAMC-BC1F03/2020|EPI_ISL_705068|20   ····················  ····················  ·····················  ····················  ····················  ····················
W   124 hCoV-19/England/CAMC-BC0405/2020|EPI_ISL_705073|20   ····················  ····················  ·····················  ····················  ····················  ····················
W   125 hCoV-19/England/CAMC-BC0BF1/2020|EPI_ISL_705076|20   ····················  ····················  ·····················  ····················  ····················  ····················
W   126 hCoV-19/England/CAMC-BC0A21/2020|EPI_ISL_705077|20   ····················  ····················  ·····················  ····················  ····················  ····················
W   127 hCoV-19/England/NORT-1B70F00/2020|EPI_ISL_706300|2   ····················  ····················  ·····················  ····················  ····················  ····················
W   128 hCoV-19/England/NORT-1B71093/2020|EPI_ISL_706301|2   ····················  ····················  ·····················  ····················  ····················  ····················
W   129 hCoV-19/England/NORT-1B710A2/2020|EPI_ISL_706302|2   ····················  ····················  ·····················  ····················  ····················  ····················
W   130 hCoV-19/England/NORT-1B71109/2020|EPI_ISL_706303|2   ····················  ····················  ·····················  ····················  ····················  ····················
W   131 hCoV-19/England/NORT-1B71163/2020|EPI_ISL_706304|2   ····················  ····················  ·····················  ····················  ····················  T···················
```

| | | | | | | | | |
|---|---|---|---|---|---|---|---|---|
| W | 132 | hCoV-19/England/NORT-1B71206/2020\|EPI_ISL_706305\|2 | ···················· | ···················· | ····················· | ···················· | ···················· | ···················· |
| W | 133 | hCoV-19/England/NORT-1B71303/2020\|EPI_ISL_706306\|2 | ···················· | ···················· | ····················· | ···················· | ···················· | ···················· |
| W | 134 | hCoV-19/England/NORT-1B71312/2020\|EPI_ISL_706307\|2 | ···················· | ···················· | ····················· | ···················· | ···················· | ···················· |
| W | 135 | hCoV-19/England/NORT-1B71321/2020\|EPI_ISL_706308\|2 | ···················· | ···················· | ····················· | ···················· | ···················· | ···················· |
| W | 136 | hCoV-19/England/NORT-1B713C7/2020\|EPI_ISL_706309\|2 | ···················· | ···················· | ····················· | ···················· | ···················· | ···················· |
| W | 137 | hCoV-19/England/NORT-1B7142E/2020\|EPI_ISL_706310\|2 | ···················· | ···················· | ····················· | ···················· | ···················· | ···················· |
| W | 138 | hCoV-19/England/NORT-1B715B2/2020\|EPI_ISL_706311\|2 | ···················· | ···················· | ····················· | ···················· | ···················· | ···················· |
| W | 139 | hCoV-19/England/NORT-1B719A7/2020\|EPI_ISL_706312\|2 | ···················· | ···················· | ····················· | ···················· | ···················· | ···················· |
| W | 140 | hCoV-19/England/NORT-1B71A95/2020\|EPI_ISL_706313\|2 | ···················· | ···················· | ····················· | ···················· | ···················· | ···················· |
| W | 141 | hCoV-19/England/NORT-1B71AA4/2020\|EPI_ISL_706314\|2 | ···················· | ···················· | ····················· | ···················· | ···················· | ···················· |
| W | 142 | hCoV-19/England/NORT-1B71AC2/2020\|EPI_ISL_706316\|2 | ···················· | ···················· | ····················· | ···················· | ···················· | ···················· |
| W | 143 | hCoV-19/England/NORT-1B71B38/2020\|EPI_ISL_706317\|2 | ···················· | ···················· | ····················· | ···················· | ···················· | ···················· |
| W | 144 | hCoV-19/England/NORT-1B71B92/2020\|EPI_ISL_706318\|2 | ···················· | ···················· | ····················· | ···················· | ···················· | ···················· |
| W | 145 | hCoV-19/England/NORT-1B71BDE/2020\|EPI_ISL_706319\|2 | ···················· | ···················· | ····················· | ···················· | ···················· | ···················· |
| W | 146 | hCoV-19/England/NORT-1B71D7E/2020\|EPI_ISL_706320\|2 | ···················· | ·A·················· | ····················· | ···················· | ···················· | ···················· |
| W | 147 | hCoV-19/England/NORT-1B71DAB/2020\|EPI_ISL_706321\|2 | ···················· | ···················· | ····················· | ···················· | ···················· | ···················· |
| W | 148 | hCoV-19/England/NORT-1B71DC9/2020\|EPI_ISL_706322\|2 | ···················· | ···················· | ····················· | ···················· | ···················· | ···················· |
| W | 149 | hCoV-19/England/NORT-1B71E02/2020\|EPI_ISL_706323\|2 | ···················· | ···················· | ····················· | ···················· | ···················· | ···················· |
| W | 150 | hCoV-19/England/NORT-1B71E3F/2020\|EPI_ISL_706324\|2 | ···················· | ···················· | ····················· | ···················· | ···················· | ···················· |
| W | 151 | hCoV-19/England/NORT-1B71EA8/2020\|EPI_ISL_706325\|2 | ···················· | ···················· | ····················· | ···················· | ···················· | ···················· |
| W | 152 | hCoV-19/England/NORT-1B71F1E/2020\|EPI_ISL_706326\|2 | ···················· | ···················· | ····················· | ···················· | ···················· | ···················· |
| W | 153 | hCoV-19/England/NORT-1B71F5A/2020\|EPI_ISL_706327\|2 | ···················· | ···················· | ····················· | ···················· | ···················· | ···················· |
| W | 154 | hCoV-19/England/NORT-1B71F96/2020\|EPI_ISL_706328\|2 | ···················· | ···················· | ····················· | ···················· | ···················· | ···················· |
| W | 155 | hCoV-19/England/NORT-1B71FA5/2020\|EPI_ISL_706329\|2 | ···················· | ···················· | ····················· | ···················· | ···················· | ···················· |
| W | 156 | hCoV-19/England/NORT-1B71FD2/2020\|EPI_ISL_706330\|2 | ···················· | ···················· | ····················· | ···················· | ···················· | ···················· |
| W | 157 | hCoV-19/England/NORT-1B71FF0/2020\|EPI_ISL_706331\|2 | ···················· | ···················· | ····················· | ···················· | ···················· | ···················· |
| W | 158 | hCoV-19/England/NORT-28D27C/2020\|EPI_ISL_706332\|20 | ···················· | ···················· | ····················· | ···················· | ···················· | ···················· |
| W | 159 | hCoV-19/England/NORT-28D29A/2020\|EPI_ISL_706333\|20 | ···················· | ·A·················· | ····················· | ···················· | ···················· | ···················· |
| W | 160 | hCoV-19/England/NORT-28D2A9/2020\|EPI_ISL_706334\|20 | ···················· | ···················· | ····················· | ···················· | ···················· | ···················· |
| W | 161 | hCoV-19/England/NORT-28D2C7/2020\|EPI_ISL_706335\|20 | ···················· | ·A·················· | ····················· | ···················· | ···················· | ···················· |
| W | 162 | hCoV-19/England/NORT-28D2D6/2020\|EPI_ISL_706336\|20 | ···················· | ···················· | ····················· | ···················· | ···················· | ···················· |
| W | 163 | hCoV-19/England/NORT-28D2F4/2020\|EPI_ISL_706337\|20 | ···················· | ···················· | ····················· | ···················· | ···················· | ···················· |
| W | 164 | hCoV-19/England/NORT-28D31F/2020\|EPI_ISL_706338\|20 | ···················· | ···················· | ····················· | ···················· | ···················· | ···················· |
| W | 165 | hCoV-19/England/NORT-28D35B/2020\|EPI_ISL_706339\|20 | ···················· | ···················· | ····················· | ···················· | ···················· | ···················· |
| W | 166 | hCoV-19/England/NORT-28D4D0/2020\|EPI_ISL_706341\|20 | ···················· | ···················· | ····················· | ···················· | ···················· | ···················· |
| W | 167 | hCoV-19/England/NORT-28D50A/2020\|EPI_ISL_706342\|20 | ···················· | ···················· | ····················· | ···················· | ···················· | ···················· |
| W | 168 | hCoV-19/England/NORT-28D519/2020\|EPI_ISL_706343\|20 | ···················· | ···················· | ····················· | ···················· | ···················· | ···················· |
| W | 169 | hCoV-19/England/NORT-28D528/2020\|EPI_ISL_706344\|20 | ···················· | ···················· | ····················· | ···················· | ···················· | ···················· |
| W | 170 | hCoV-19/England/NORT-28D546/2020\|EPI_ISL_706345\|20 | ···················· | ···················· | ····················· | ···················· | ···················· | ···················· |
| W | 171 | hCoV-19/England/NORT-28D555/2020\|EPI_ISL_706346\|20 | ···················· | ···················· | ····················· | ···················· | ···················· | ···················· |
| W | 172 | hCoV-19/England/NORT-28D564/2020\|EPI_ISL_706347\|20 | ···················· | ···················· | ····················· | ···················· | ···················· | ···················· |
| W | 173 | hCoV-19/England/NORT-28D573/2020\|EPI_ISL_706348\|20 | ···················· | ···················· | ····················· | ···················· | ···················· | ···················· |
| W | 174 | hCoV-19/England/NORT-28D582/2020\|EPI_ISL_706349\|20 | ···················· | ···················· | ····················· | ···················· | ···················· | ···················· |
| W | 175 | hCoV-19/England/NORT-28D607/2020\|EPI_ISL_706350\|20 | ···················· | ···················· | ····················· | ···················· | ····T··············· | ···················· |
| W | 176 | hCoV-19/England/NORT-28D69E/2020\|EPI_ISL_706351\|20 | ···················· | ···················· | ····················· | ···················· | ···················· | ···················· |
| W | 177 | hCoV-19/England/NORT-28D6BC/2020\|EPI_ISL_706352\|20 | ···················· | ···················· | ····················· | ···················· | ···················· | ···················· |
| W | 178 | hCoV-19/England/NORT-2A7467/2020\|EPI_ISL_706353\|20 | ···················· | ···················· | ····················· | ···················· | ···················· | ···················· |
| W | 179 | hCoV-19/England/NORT-2A75A0/2020\|EPI_ISL_706354\|20 | ···················· | ···················· | ····················· | ···················· | ···················· | ···················· |
| W | 180 | hCoV-19/England/NORW-EED4D/2020\|EPI_ISL_706356\|202 | ···················· | ···················· | ····················· | ···················· | ···················· | ···················· |
| W | 181 | hCoV-19/England/NORW-EED6B/2020\|EPI_ISL_706357\|202 | ···················· | ···················· | ····················· | ···················· | ···················· | ···················· |
| W | 182 | hCoV-19/England/NORW-EEDA7/2020\|EPI_ISL_706358\|202 | ···················· | ···················· | ····················· | ···················· | ···················· | ···················· |
| W | 183 | hCoV-19/England/NORW-EEDE3/2020\|EPI_ISL_706359\|202 | ···················· | ···················· | ····················· | ···················· | ···················· | ···················· |
| W | 184 | hCoV-19/England/NORW-EEEB3/2020\|EPI_ISL_706360\|202 | ···················· | ···················· | ····················· | ···················· | ···················· | ···················· |
| W | 185 | hCoV-19/England/NORW-F6207/2020\|EPI_ISL_706361\|202 | ···················· | ·A·················· | ····················· | ···················· | ···················· | ···················· |
| W | 187 | hCoV-19/England/NORW-F62F8/2020\|EPI_ISL_706363\|202 | ···················· | ···················· | ····················· | ···················· | ···················· | ···················· |
| W | 188 | hCoV-19/England/NORW-F639B/2020\|EPI_ISL_706364\|202 | ···················· | ···················· | ····················· | ···················· | ···················· | ···················· |
| W | 189 | hCoV-19/England/NORW-F63F5/2020\|EPI_ISL_706365\|202 | ···················· | ···················· | ····················· | ···················· | ···················· | ···················· |
| W | 190 | hCoV-19/England/NORW-F646B/2020\|EPI_ISL_706366\|202 | ···················· | ···················· | ····················· | ···················· | ···················· | ···················· |
| W | 191 | hCoV-19/England/NORW-F653B/2020\|EPI_ISL_706367\|202 | ···················· | ···················· | ····················· | ···················· | ···················· | ···················· |
| W | 192 | hCoV-19/England/NORW-F6568/2020\|EPI_ISL_706368\|202 | ···················· | ···················· | ····················· | ···················· | ···················· | ···················· |
| W | 193 | hCoV-19/England/NORW-F65B3/2020\|EPI_ISL_706369\|202 | ···················· | ···················· | ····················· | ···················· | ···················· | ···················· |
| W | 194 | hCoV-19/England/NORW-F65D1/2020\|EPI_ISL_706370\|202 | ···················· | ···················· | ····················· | ···················· | ···················· | ···················· |
| W | 195 | hCoV-19/England/NORW-F65E0/2020\|EPI_ISL_706371\|202 | ···················· | ···················· | ····················· | ···················· | ···················· | ···················· |
| W | 196 | hCoV-19/England/NORW-F6629/2020\|EPI_ISL_706372\|202 | ···················· | ···················· | ····················· | ···················· | ···················· | ···················· |
| W | 197 | hCoV-19/England/NORW-F6647/2020\|EPI_ISL_706373\|202 | ···················· | ···················· | ····················· | ···················· | ···················· | ···················· |
| W | 198 | hCoV-19/England/NORW-F6717/2020\|EPI_ISL_706374\|202 | ···················· | ···················· | ····················· | ···················· | ···················· | ···················· |
| W | 199 | hCoV-19/England/NORW-F6726/2020\|EPI_ISL_706375\|202 | ···················· | ···················· | ····················· | ···················· | ···················· | ···················· |
| W | 200 | hCoV-19/England/NORW-F6744/2020\|EPI_ISL_706376\|202 | ···················· | ···················· | ····················· | ···················· | ···················· | ···················· |
| W | 201 | hCoV-19/England/NORW-F6771/2020\|EPI_ISL_706377\|202 | ···················· | ···················· | ····················· | ···················· | ···················· | ···················· |
| W | 202 | hCoV-19/England/NORW-F6B66/2020\|EPI_ISL_706378\|202 | ···················· | ···················· | ····················· | ···················· | ···················· | ···················· |
| W | 203 | hCoV-19/England/NORW-F6BDF/2020\|EPI_ISL_706379\|202 | ···················· | ···················· | ····················· | ···················· | ···················· | ···················· |
| W | 204 | hCoV-19/England/NORW-F6C18/2020\|EPI_ISL_706380\|202 | ···················· | ···················· | ····················· | ···················· | ···················· | ···················· |
| W | 205 | hCoV-19/England/NORW-F6C36/2020\|EPI_ISL_706381\|202 | ···················· | ···················· | ····················· | ···················· | ···················· | ···················· |
| W | 206 | hCoV-19/England/NORW-F6C72/2020\|EPI_ISL_706382\|202 | ···················· | ···················· | ····················· | ···················· | ···················· | ···················· |
| W | 207 | hCoV-19/England/NORW-F6F88/2020\|EPI_ISL_706386\|202 | ···················· | ···················· | ····················· | ···················· | ···················· | ···················· |
| W | 208 | hCoV-19/England/NORW-F6FF1/2020\|EPI_ISL_706387\|202 | ···················· | ···················· | ····················· | ···················· | ···················· | ···················· |
| W | 209 | hCoV-19/England/NORW-F7172/2020\|EPI_ISL_706390\|202 | ···················· | ···················· | ····················· | ···················· | ···················· | ···················· |
| W | 210 | hCoV-19/England/NORW-F7251/2020\|EPI_ISL_706391\|202 | ···················· | ···················· | ····················· | ···················· | ···················· | ···················· |
| W | 211 | hCoV-19/England/NORW-F734F/2020\|EPI_ISL_706392\|202 | ···················· | ···················· | ····················· | ···················· | ···················· | ···················· |
| W | 212 | hCoV-19/England/NORW-F737C/2020\|EPI_ISL_706393\|202 | ···················· | ···················· | ····················· | ···················· | ···················· | ···················· |

```
W 213 hCoV-19/England/NORW-F73A9/2020|EPI_ISL_706394|202 ···················· ···················· ···················· ···················· ···················· ····················
W 214 hCoV-19/England/NORW-F73D6/2020|EPI_ISL_706395|202 ···················· ···················· ···················· ···················· ···················· ····················
W 215 hCoV-19/England/NORW-F753A/2020|EPI_ISL_706396|202 ···················· ···················· ···················· ···················· ···················· ····················
W 216 hCoV-19/England/NORW-F75C1/2020|EPI_ISL_706397|202 ···················· ···················· ···················· ···················· ···················· ····················
W 217 hCoV-19/England/NORW-F75D0/2020|EPI_ISL_706398|202 ···················· ···················· ···················· ···················· ···················· ····················
W 218 hCoV-19/England/NORW-F7619/2020|EPI_ISL_706399|202 ···················· ···················· ···················· ···················· ···················· ····················
W 219 hCoV-19/Indonesia/BT-SHSIU-01-3610/2020|EPI_ISL_70 ···················· ···A················ ···················· ···················· ···················· ····················
W 220 hCoV-19/USA/CA-LACPHL-AD00003/2020|EPI_ISL_707693| ···················· ···················· ···················· ···················· ···················· ····················
W 221 hCoV-19/USA/CA-LACPHL-AB00002/2020|EPI_ISL_707694| ···················· ···················· ···················· ···················· ···················· ····················
W 222 hCoV-19/USA/CA-LACPHL-AB00003/2020|EPI_ISL_707695| ···················· ···················· ···················· ···················· ···················· ····················
W 223 hCoV-19/Indonesia/BT-SHSIU-01-4941/2020|EPI_ISL_70 ···················· ···················· ···················· ···················· ···················· ····················
W 224 hCoV-19/Tunisia/61628/2020|EPI_ISL_707697|2020-11- ···················· ···················· ···················· ···················· ···················· ····················
W 225 hCoV-19/Tunisia/61627/2020|EPI_ISL_707698|2020-11- ···················· ···················· ···················· ···················· ···················· ····················
W 226 hCoV-19/Tunisia/61624/2020|EPI_ISL_707699|2020-11- ···················· ···················· ···················· ···················· ···················· ····················
W 227 hCoV-19/England/ALDP-BE957C/2020|EPI_ISL_708950|20 ···················· ···················· ···················· ···················· ···················· ····················
W 228 hCoV-19/England/MILK-BEE243/2020|EPI_ISL_708951|20 ···················· ···················· ···················· ···················· ···················· ····················
W 229 hCoV-19/England/QEUH-BEB01F/2020|EPI_ISL_708953|20 ···················· ···················· ···················· ···················· ···················· ····················
W 230 hCoV-19/England/ALDP-BE9345/2020|EPI_ISL_708955|20 ···················· ···················· ···················· ···················· ···················· ····················
W 231 hCoV-19/England/ALDP-BE8F8E/2020|EPI_ISL_708956|20 ···················· ···················· ···················· ···················· ···················· ····················
W 232 hCoV-19/England/QEUH-BEB03D/2020|EPI_ISL_708957|20 ···················· ···················· ···················· ···················· ···················· ····················
W 233 hCoV-19/England/ALDP-BE966A/2020|EPI_ISL_708959|20 ···················· ·A·················· ···················· ···················· ···················· ··A·················
W 234 hCoV-19/England/ALDP-BE9354/2020|EPI_ISL_708960|20 ···················· ···················· ···················· ···················· ···················· ····················
W 235 hCoV-19/England/ALDP-BE97D0/2020|EPI_ISL_708961|20 ···················· ···················· ···················· ···················· ···················· ····················
W 236 hCoV-19/England/ALDP-BE9688/2020|EPI_ISL_708962|20 ···················· ···················· ···················· ···················· ···················· ····················
W 237 hCoV-19/England/ALDP-BE8F9D/2020|EPI_ISL_708967|20 ···················· ···················· ···················· ···················· ···················· ····················
W 238 hCoV-19/England/ALDP-BE8FCA/2020|EPI_ISL_708968|20 ···················· ···················· ···················· ···················· ···················· ····················
W 239 hCoV-19/England/ALDP-BE910F/2020|EPI_ISL_708972|20 ···················· ···················· ···················· ···················· ···················· ····················
W 240 hCoV-19/England/ALDP-BE8E36/2020|EPI_ISL_708973|20 ···················· ···················· ···················· ···················· ···················· ····················
W 241 hCoV-19/England/ALDP-BE905D/2020|EPI_ISL_708974|20 ···················· ···················· ···················· ···················· ···················· ····················
W 242 hCoV-19/England/ALDP-BE91D2/2020|EPI_ISL_708976|20 ···················· ···················· ···················· ···················· ···················· ····················
W 243 hCoV-19/England/ALDP-BE9697/2020|EPI_ISL_708977|20 ···················· ···················· ···················· ···················· ···················· ····················
W 244 hCoV-19/England/ALDP-BE956D/2020|EPI_ISL_708978|20 ···················· ···················· ···················· ···················· ···················· ····················
W 245 hCoV-19/England/MILK-BEE38C/2020|EPI_ISL_708979|20 ···················· ···················· ···················· ···················· ···················· ····················
W 246 hCoV-19/England/ALDP-BE921B/2020|EPI_ISL_708980|20 ···················· ···················· ···················· ···················· ···················· ····················
W 247 hCoV-19/England/ALDP-BE8EDC/2020|EPI_ISL_708981|20 ···················· ···················· ···················· ···················· ·····T·············· ····················
W 248 hCoV-19/England/ALDP-BE93CD/2020|EPI_ISL_708982|20 ···················· ···················· ···················· ···················· ···················· ····················
W 249 hCoV-19/England/MILK-BEE4C5/2020|EPI_ISL_708983|20 ···················· ···················· ···················· ···················· ···················· ····················
W 250 hCoV-19/England/MILK-BEE36E/2020|EPI_ISL_708984|20 ···················· ···················· ···················· ···················· ···················· ····················
W 251 hCoV-19/England/QEUH-BEB079/2020|EPI_ISL_708985|20 ···················· ···················· ···················· ···················· ···················· ····················
W 252 hCoV-19/England/MILK-BEE225/2020|EPI_ISL_708986|20 ···················· ···················· ···················· ···················· ···················· ····················
W 253 hCoV-19/England/ALDP-BE8F60/2020|EPI_ISL_708987|20 ···················· ···················· ···················· ···················· ···················· ····················
W 254 hCoV-19/England/ALDP-BE8F06/2020|EPI_ISL_708988|20 ···················· ···················· ···················· ···················· ···················· ····················
W 255 hCoV-19/England/CAMC-BFB850/2020|EPI_ISL_708992|20 ···················· ···················· ···················· ···················· ···················· ····················
W 256 hCoV-19/England/CAMC-BFBFE2/2020|EPI_ISL_708996|20 ···················· ···················· ···················· ···················· ···················· ····················
W 257 hCoV-19/USA/AZ-TG475423/2020|EPI_ISL_695049|2020-0 ···················· ···················· ···················· ···················· ···················· ····················
W 258 hCoV-19/USA/AZ-TG478871/2020|EPI_ISL_695053|2020-0 ···················· ···················· ···················· ···················· ···················· ····················
W 259 hCoV-19/USA/AZ-TG479330/2020|EPI_ISL_695057|2020-0 ···················· ···················· ···················· ···················· ···················· ····················
W 260 hCoV-19/USA/AZ-TG480043/2020|EPI_ISL_695061|2020-0 ···················· ·A·················· ···················· ···················· ···················· ····················
W 261 hCoV-19/USA/AZ-TG480342/2020|EPI_ISL_695065|2020-0 ···················· ···················· ···················· ···················· ···················· ····················
W 262 hCoV-19/USA/AZ-TG481285/2020|EPI_ISL_695073|2020-0 ···················· ···················· ···················· ···················· ···················· ····················
W 263 hCoV-19/USA/AZ-TG482920/2020|EPI_ISL_695074|2020-0 ···················· ···················· ···················· ···················· ···················· ····················
W 264 hCoV-19/USA/AZ-TG483179/2020|EPI_ISL_695075|2020-0 ···················· ·A·················· ···················· ···················· ···················· ····················
W 265 hCoV-19/USA/AZ-TG483180/2020|EPI_ISL_695076|2020-0 ···················· ·A·················· ···················· ···················· ···················· ····················
W 266 hCoV-19/USA/AZ-TG487006/2020|EPI_ISL_695093|2020-0 ···················· ···················· ···················· ···················· ···················· ····················
W 267 hCoV-19/USA/AZ-TG668394/2020|EPI_ISL_696300|2020-0 ···················· ···················· ···················· ···················· ···················· ····················
W 268 hCoV-19/USA/AZ-TG668395/2020|EPI_ISL_696301|2020-0 ···················· ···················· ···················· ···················· ···················· ····················
W 269 hCoV-19/USA/AZ-TG668396/2020|EPI_ISL_696302|2020-0 ···················· ···················· ···················· ···················· ···················· ····················
W 270 hCoV-19/USA/AZ-TG668398/2020|EPI_ISL_696303|2020-0 ···················· ···················· ···················· ···················· ···················· ····················
W 271 hCoV-19/USA/AZ-TG668400/2020|EPI_ISL_696304|2020-0 ···················· ···················· ···················· ···················· ···················· ····················
W 272 hCoV-19/USA/AZ-TG668409/2020|EPI_ISL_696310|2020-0 ···················· ···················· ···················· ···················· ···················· ····················
W 273 hCoV-19/USA/AZ-TG668418/2020|EPI_ISL_696313|2020-0 ···················· ···················· ···················· ···················· ···················· ····················
W 274 hCoV-19/USA/AZ-TG670093/2020|EPI_ISL_696316|2020-0 ···················· ···················· ···················· ···················· ···················· ····················
W 275 hCoV-19/USA/AZ-TG670096/2020|EPI_ISL_696318|2020-0 ···················· ···················· ···················· ···················· ···················· ····················
W 276 hCoV-19/USA/AZ-TG670098/2020|EPI_ISL_696320|2020-0 ···················· ···················· ···················· ···················· ···················· ····················
W 277 hCoV-19/USA/AZ-TG670099/2020|EPI_ISL_696321|2020-0 ···················· ···················· ···················· ···················· ···················· ····················
W 278 hCoV-19/USA/AZ-TG670189/2020|EPI_ISL_696384|2020-0 ···················· ···················· ···················· ···················· ···················· ····················
W 279 hCoV-19/USA/AZ-TG670201/2020|EPI_ISL_696392|2020-0 ···················· ···················· ···················· ···················· ···················· ····················
W 280 hCoV-19/USA/AZ-TG670204/2020|EPI_ISL_696394|2020-0 ···················· ···················· ···················· ···················· ···················· ····················
W 281 hCoV-19/USA/AZ-TG670207/2020|EPI_ISL_696397|2020-0 ···················· ···················· ···················· ···················· ···················· ····················
W 282 hCoV-19/USA/AZ-TG670224/2020|EPI_ISL_696407|2020-0 ···················· ···················· ···················· ···················· ···················· ····················
W 283 hCoV-19/USA/AZ-TG670226/2020|EPI_ISL_696409|2020-0 ···················· ···················· ···················· ···················· ···················· ····················
W 284 hCoV-19/USA/AZ-TG670228/2020|EPI_ISL_696410|2020-0 ···················· ···················· ···················· ···················· ···················· ····················
W 285 hCoV-19/United Arab Emirates/0814/2020|EPI_ISL_698 ···················· ···················· ···················· ···················· ···················· ····················
W 286 hCoV-19/United Arab Emirates/0815/2020|EPI_ISL_698 ···················· ···················· ···················· ···················· ···················· ····················
W 287 hCoV-19/United Arab Emirates/0816/2020|EPI_ISL_698 ···················· ···················· ···················· ···················· ···················· ····················
W 288 hCoV-19/United Arab Emirates/0817/2020|EPI_ISL_698 ···················· ···················· ···················· ···················· ···················· ····················
W 289 hCoV-19/United Arab Emirates/0818/2020|EPI_ISL_698 ···················· ···················· ···················· ···················· ···················· ····················
W 290 hCoV-19/United Arab Emirates/0819/2020|EPI_ISL_698 ···················· ···················· ···················· ···················· ···················· ····················
W 291 hCoV-19/United Arab Emirates/0820/2020|EPI_ISL_698 ···················· ···················· ···················· ···················· ···················· ····················
W 292 hCoV-19/United Arab Emirates/0821/2020|EPI_ISL_698 ···················· ···················· ···················· ···················· ···················· ····················
```

| | | | | | | | |
|---|---|---|---|---|---|---|---|
| W | 293 hCoV-19/United Arab Emirates/0822/2020\|EPI_ISL_698 | ···················· | ···················· | ···················· | ···················· | ···················· | ···················· |
| W | 294 hCoV-19/United Arab Emirates/0823/2020\|EPI_ISL_698 | ···················· | ···················· | ···················· | ···················· | ···················· | ···················· |
| W | 295 hCoV-19/United Arab Emirates/0824/2020\|EPI_ISL_698 | ···················· | ···················· | ···················· | ···················· | ···················· | ···················· |
| W | 296 hCoV-19/United Arab Emirates/0825/2020\|EPI_ISL_698 | ···················· | ···················· | ···················· | ···················· | ···················· | ···················· |
| W | 297 hCoV-19/United Arab Emirates/0826/2020\|EPI_ISL_698 | ···················· | ···················· | ···················· | ···················· | ···················· | ···················· |
| W | 298 hCoV-19/United Arab Emirates/0827/2020\|EPI_ISL_698 | ···················· | ···················· | ···················· | ···················· | ···················· | ···················· |
| W | 299 hCoV-19/United Arab Emirates/0828/2020\|EPI_ISL_698 | ···················· | ···················· | ···················· | ···················· | ···················· | ···················· |
| W | 300 hCoV-19/United Arab Emirates/0829/2020\|EPI_ISL_698 | ···················· | ···················· | ···················· | ···················· | ···················· | ···················· |
| W | 301 hCoV-19/United Arab Emirates/0830/2020\|EPI_ISL_698 | ···················· | ···················· | ···················· | ···················· | ···················· | ···················· |
| W | 302 hCoV-19/United Arab Emirates/0831/2020\|EPI_ISL_698 | ···················· | ···················· | ···················· | ···················· | ···················· | ···················· |
| W | 303 hCoV-19/United Arab Emirates/0832/2020\|EPI_ISL_698 | ···················· | ···················· | ···················· | ···················· | ···················· | ···················· |
| W | 304 hCoV-19/United Arab Emirates/0833/2020\|EPI_ISL_698 | ···················· | ···················· | ···················· | ···················· | ···················· | ···················· |
| W | 305 hCoV-19/United Arab Emirates/0834/2020\|EPI_ISL_698 | ···················· | ···················· | ···················· | ···················· | ···················· | ···················· |
| W | 306 hCoV-19/United Arab Emirates/0835/2020\|EPI_ISL_698 | ···················· | ···················· | ···················· | ···················· | ···················· | ···················· |
| W | 307 hCoV-19/United Arab Emirates/0836/2020\|EPI_ISL_698 | ···················· | ···················· | ···················· | ···················· | ···················· | ···················· |
| W | 308 hCoV-19/United Arab Emirates/0837/2020\|EPI_ISL_698 | ···················· | ···················· | ···················· | ···················· | ···················· | ···················· |
| W | 309 hCoV-19/United Arab Emirates/0838/2020\|EPI_ISL_698 | ···················· | ···················· | ···················· | ···················· | ···················· | ···················· |
| W | 310 hCoV-19/United Arab Emirates/0839/2020\|EPI_ISL_698 | ···················· | ···················· | ···················· | ···················· | ···················· | ···················· |
| W | 311 hCoV-19/United Arab Emirates/0840/2020\|EPI_ISL_698 | ···················· | ···················· | ···················· | ···················· | ···················· | ···················· |
| W | 312 hCoV-19/United Arab Emirates/0843/2020\|EPI_ISL_698 | ···················· | ···················· | ···················· | ···················· | ···················· | ···················· |
| W | 313 hCoV-19/United Arab Emirates/0844/2020\|EPI_ISL_698 | ···················· | ···················· | ···················· | ···················· | ···················· | ···················· |
| W | 314 hCoV-19/United Arab Emirates/0845/2020\|EPI_ISL_698 | ···················· | ···················· | ···················· | ···················· | ···················· | ···················· |
| W | 315 hCoV-19/United Arab Emirates/0846/2020\|EPI_ISL_698 | ···················· | ···················· | ···················· | ···················· | ···················· | ···················· |
| W | 316 hCoV-19/India/MH-ACTREC-555/2020\|EPI_ISL_700200\|20 | ···················· | ···················· | ···················· | ···················· | ···················· | ···················· |
| W | 317 hCoV-19/India/MH-ACTREC-556/2020\|EPI_ISL_700201\|20 | ···················· | ···················· | ···················· | ···················· | ···················· | ···················· |
| W | 318 hCoV-19/India/MH-ACTREC-557/2020\|EPI_ISL_700202\|20 | ···················· | ···················· | ···················· | ···················· | ···················· | ···················· |
| W | 320 hCoV-19/India/MH-ACTREC-559/2020\|EPI_ISL_700204\|20 | ···················· | ···················· | ···················· | ···················· | ···················· | ···················· |
| W | 321 hCoV-19/India/MH-ACTREC-560/2020\|EPI_ISL_700205\|20 | ···················· | ···················· | ···················· | ···················· | ···················· | ···················· |
| W | 322 hCoV-19/India/MH-ACTREC-561/2020\|EPI_ISL_700206\|20 | ···················· | ···················· | ···················· | ···················· | ···················· | ···················· |
| W | 323 hCoV-19/India/MH-ACTREC-562/2020\|EPI_ISL_700207\|20 | ···················· | ···················· | ···················· | ···················· | ···················· | ···················· |
| W | 324 hCoV-19/India/MH-ACTREC-563/2020\|EPI_ISL_700208\|20 | ···················· | ···················· | ···················· | ···················· | ···················· | ···················· |
| W | 326 hCoV-19/India/MH-ACTREC-565/2020\|EPI_ISL_700210\|20 | ···················· | ···················· | ···················· | ···················· | ···················· | ···················· |
| W | 327 hCoV-19/India/MH-ACTREC-567/2020\|EPI_ISL_700211\|20 | ···················· | ···················· | ···················· | ···················· | ···················· | ···················· |
| W | 328 hCoV-19/India/MH-ACTREC-568/2020\|EPI_ISL_700212\|20 | ···················· | ···················· | ···················· | ···················· | ···················· | ···················· |
| W | 329 hCoV-19/India/MH-ACTREC-570/2020\|EPI_ISL_700214\|20 | ···················· | ···················· | ···················· | ···················· | ···················· | ···················· |
| W | 330 hCoV-19/India/MH-ACTREC-571/2020\|EPI_ISL_700215\|20 | ···················· | ···················· | ···················· | ···················· | ···················· | ···················· |
| W | 332 hCoV-19/India/MH-ACTREC-573/2020\|EPI_ISL_700217\|20 | ···················· | ···················· | ···················· | ···················· | ···················· | ···················· |
| W | 333 hCoV-19/India/MH-ACTREC-574/2020\|EPI_ISL_700218\|20 | ···················· | ·A·················· | ···················· | ···················· | ···················· | ···················· |
| W | 335 hCoV-19/India/MH-ACTREC-578/2020\|EPI_ISL_700221\|20 | ···················· | ···················· | ···················· | ···················· | ···················· | ···················· |
| W | 336 hCoV-19/India/MH-ACTREC-580/2020\|EPI_ISL_700222\|20 | ···················· | ·A·················· | ···················· | ···················· | ···················· | ···················· |
| W | 337 hCoV-19/India/MH-ACTREC-582/2020\|EPI_ISL_700223\|20 | ···················· | ···················· | ···················· | ···················· | ···················· | ···················· |
| W | 338 hCoV-19/India/MH-ACTREC-585/2020\|EPI_ISL_700224\|20 | ···················· | ···················· | ···················· | ···················· | ···················· | ···················· |
| W | 339 hCoV-19/India/MH-ACTREC-586/2020\|EPI_ISL_700225\|20 | ···················· | ···················· | ···················· | ···················· | ···················· | ···················· |
| W | 342 hCoV-19/India/MH-ACTREC-590/2020\|EPI_ISL_700228\|20 | ···················· | ···················· | ···················· | ···················· | ···················· | ···················· |
| W | 343 hCoV-19/India/MH-ACTREC-591/2020\|EPI_ISL_700229\|20 | ···················· | ·A·················· | ···················· | ···················· | ···················· | ···A················ |
| W | 345 hCoV-19/India/MH-ACTREC-593/2020\|EPI_ISL_700231\|20 | ···················· | ·A·················· | ···················· | ···················· | ···················· | ···················· |
| W | 346 hCoV-19/India/MH-ACTREC-594/2020\|EPI_ISL_700232\|20 | ···················· | ·A·················· | ···················· | ···················· | ···················· | ···················· |
| W | 347 hCoV-19/India/MH-ACTREC-595/2020\|EPI_ISL_700233\|20 | ···················· | ·A·················· | ···················· | ···················· | ···················· | ···A················ |
| W | 348 hCoV-19/India/MH-ACTREC-596/2020\|EPI_ISL_700234\|20 | ···················· | ·A·················· | ···················· | ···················· | ···················· | ···················· |
| W | 350 hCoV-19/India/MH-ACTREC-598/2020\|EPI_ISL_700236\|20 | ···················· | ···················· | ···················· | ···················· | ···················· | ···················· |
| W | 351 hCoV-19/India/MH-ACTREC-599/2020\|EPI_ISL_700237\|20 | ···················· | ···················· | ···················· | ···················· | ···················· | ···················· |
| W | 352 hCoV-19/India/MH-ACTREC-600/2020\|EPI_ISL_700238\|20 | ···················· | ···················· | ···················· | ···················· | ···················· | ···················· |
| W | 353 hCoV-19/India/MH-ACTREC-601/2020\|EPI_ISL_700239\|20 | ···················· | ···················· | ···················· | ···················· | ···················· | ···················· |
| W | 355 hCoV-19/India/MH-ACTREC-604/2020\|EPI_ISL_700241\|20 | ···················· | ·A·················· | ···················· | ···················· | ···················· | ···A················ |
| W | 358 hCoV-19/India/MH-ACTREC-607/2020\|EPI_ISL_700244\|20 | ···················· | ···················· | ···················· | ···················· | ···················· | ···················· |
| W | 359 hCoV-19/India/MH-ACTREC-608/2020\|EPI_ISL_700245\|20 | ···················· | ···················· | ···················· | ···················· | ···················· | ···················· |
| W | 360 hCoV-19/India/MH-ACTREC-609/2020\|EPI_ISL_700246\|20 | ···················· | ···················· | ···················· | ···················· | ···················· | ···················· |
| W | 361 hCoV-19/India/MH-ACTREC-610/2020\|EPI_ISL_700247\|20 | ···················· | ···················· | ···················· | ···················· | ···················· | ···················· |
| W | 362 hCoV-19/India/MH-ACTREC-611/2020\|EPI_ISL_700248\|20 | ···················· | ···················· | ···················· | ···················· | ···················· | ···················· |
| W | 363 hCoV-19/India/MH-ACTREC-612/2020\|EPI_ISL_700249\|20 | ···················· | ···················· | ···················· | ···················· | ···················· | ···················· |
| W | 364 hCoV-19/India/MH-ACTREC-613/2020\|EPI_ISL_700250\|20 | ···················· | ···················· | ···················· | ···················· | ···················· | ···················· |
| W | 365 hCoV-19/India/MH-ACTREC-616/2020\|EPI_ISL_700251\|20 | ···················· | ···················· | ···················· | ···················· | ···················· | ···················· |
| W | 366 hCoV-19/India/MH-ACTREC-617/2020\|EPI_ISL_700252\|20 | ···················· | ···················· | ···················· | ···················· | ···················· | ···················· |
| W | 368 hCoV-19/India/MH-ACTREC-619/2020\|EPI_ISL_700254\|20 | ···················· | ···················· | ···················· | ···················· | ···················· | ···················· |
| W | 369 hCoV-19/India/MH-ACTREC-620/2020\|EPI_ISL_700255\|20 | ···················· | ···················· | ···················· | ···················· | ···················· | ···················· |
| W | 370 hCoV-19/India/MH-ACTREC-621/2020\|EPI_ISL_700256\|20 | ···················· | ···················· | ···················· | ···················· | ···················· | ···················· |
| W | 371 hCoV-19/India/MH-ACTREC-622/2020\|EPI_ISL_700257\|20 | ···················· | ···················· | ···················· | ···················· | ···················· | ···················· |
| W | 372 hCoV-19/India/MH-ACTREC-623/2020\|EPI_ISL_700258\|20 | ···················· | ···················· | ···················· | ···················· | ···················· | ···················· |
| W | 373 hCoV-19/India/MH-ACTREC-624/2020\|EPI_ISL_700259\|20 | ···················· | ···················· | ···················· | ···················· | ···················· | ···················· |
| W | 374 hCoV-19/India/MH-ACTREC-625/2020\|EPI_ISL_700260\|20 | ···················· | ·A·················· | ···················· | ···················· | ···················· | ···A················ |
| W | 375 hCoV-19/India/MH-ACTREC-626/2020\|EPI_ISL_700261\|20 | ···················· | ·A·················· | ···················· | ···················· | ···················· | ···A················ |
| W | 376 hCoV-19/India/MH-ACTREC-627/2020\|EPI_ISL_700262\|20 | ···················· | ···················· | ···················· | ···················· | ···················· | ···················· |
| W | 378 hCoV-19/India/MH-ACTREC-629/2020\|EPI_ISL_700264\|20 | ···················· | ······A············· | ···················· | ···················· | ···················· | ···················· |
| W | 379 hCoV-19/India/MH-ACTREC-632/2020\|EPI_ISL_700265\|20 | ···················· | ···················· | ···················· | ···················· | ···················· | ···················· |
| W | 380 hCoV-19/India/MH-ACTREC-633/2020\|EPI_ISL_700266\|20 | ···················· | ···················· | ···················· | ···················· | ···················· | ···················· |
| W | 382 hCoV-19/India/MH-ACTREC-635/2020\|EPI_ISL_700268\|20 | ···················· | ·A·················· | ···················· | ···················· | ···················· | ···················· |
| W | 383 hCoV-19/India/MH-ACTREC-636/2020\|EPI_ISL_700269\|20 | ···················· | ···················· | ···················· | ···················· | ···················· | ···················· |
| W | 384 hCoV-19/India/MH-ACTREC-637/2020\|EPI_ISL_700270\|20 | ···················· | ···················· | ···················· | ···················· | ···················· | ···················· |
| W | 385 hCoV-19/India/MH-ACTREC-638/2020\|EPI_ISL_700271\|20 | ···················· | ···················· | ···················· | ···················· | ···················· | ···················· |
| W | 387 hCoV-19/India/MH-ACTREC-640/2020\|EPI_ISL_700273\|20 | ···················· | ·A·················· | ···················· | ···················· | ···················· | ···A················ |

| | | | | | | | |
|---|---|---|---|---|---|---|---|
| W | 388 hCoV-19/India/MH-ACTREC-641/2020\|EPI_ISL_700274\|20 | ···················· | ·A·················· | ···················· | ···················· | ···················· | ··A················· |
| W | 389 hCoV-19/India/MH-ACTREC-642/2020\|EPI_ISL_700275\|20 | ···················· | ·A·················· | ···················· | ···················· | ···················· | ··A················· |
| W | 390 hCoV-19/India/MH-ACTREC-646/2020\|EPI_ISL_700276\|20 | ···················· | ·A·················· | ···················· | ···················· | ···················· | ··A················· |
| W | 391 hCoV-19/India/MH-ACTREC-647/2020\|EPI_ISL_700277\|20 | ···················· | ·A·················· | ···················· | ···················· | ···················· | ··A················· |
| W | 392 hCoV-19/India/MH-ACTREC-648/2020\|EPI_ISL_700278\|20 | ···················· | ···················· | ···················· | ···················· | ···················· | ···················· |
| W | 393 hCoV-19/India/MH-ACTREC-649/2020\|EPI_ISL_700279\|20 | ···················· | ···················· | ···················· | ···················· | ···················· | ···················· |
| W | 394 hCoV-19/India/MH-ACTREC-650/2020\|EPI_ISL_700280\|20 | ···················· | ···················· | ···················· | ···················· | ···················· | ···················· |
| W | 395 hCoV-19/India/MH-ACTREC-651/2020\|EPI_ISL_700281\|20 | ···················· | ···················· | ···················· | ···················· | ···················· | ···················· |
| W | 396 hCoV-19/India/MH-ACTREC-652/2020\|EPI_ISL_700282\|20 | ···················· | ···················· | ···················· | ···················· | ···················· | ···················· |
| W | 397 hCoV-19/India/MH-ACTREC-653/2020\|EPI_ISL_700283\|20 | ···················· | ···················· | ···················· | ···················· | ···················· | ···················· |
| W | 398 hCoV-19/India/MH-ACTREC-654/2020\|EPI_ISL_700284\|20 | ···················· | ···················· | ···················· | ···················· | ···················· | ···················· |
| W | 399 hCoV-19/India/MH-ACTREC-655/2020\|EPI_ISL_700285\|20 | ···················· | ···················· | ···················· | ···················· | ···················· | ···················· |
| W | 400 hCoV-19/India/MH-ACTREC-656/2020\|EPI_ISL_700286\|20 | ···················· | ···················· | ···················· | ···················· | ···················· | ···················· |
| W | 401 hCoV-19/India/MH-ACTREC-657/2020\|EPI_ISL_700287\|20 | ···················· | ···················· | ···················· | ···················· | ···················· | ···················· |
| W | 402 hCoV-19/India/MH-ACTREC-658/2020\|EPI_ISL_700288\|20 | ···················· | ···················· | ···················· | ···················· | ···················· | ··R················· |
| W | 403 hCoV-19/India/MH-ACTREC-659/2020\|EPI_ISL_700289\|20 | ···················· | ·A·················· | ···················· | ···················· | ···················· | ··A················· |
| W | 404 hCoV-19/India/MH-ACTREC-660/2020\|EPI_ISL_700290\|20 | ···················· | ···················· | ···················· | ···················· | ···················· | ···················· |
| W | 405 hCoV-19/India/MH-ACTREC-662/2020\|EPI_ISL_700291\|20 | ···················· | ···················· | ···················· | ···················· | ···················· | ···················· |
| W | 406 hCoV-19/India/MH-ACTREC-663/2020\|EPI_ISL_700292\|20 | ···················· | ···················· | ···················· | ···················· | ···················· | ···················· |
| W | 407 hCoV-19/India/MH-ACTREC-664/2020\|EPI_ISL_700293\|20 | ···················· | ·A·················· | ···················· | ···················· | ·····K·············· | ··A················· |
| W | 408 hCoV-19/India/MH-ACTREC-668/2020\|EPI_ISL_700294\|20 | ···················· | ···················· | ···················· | ···················· | ···················· | ···················· |
| W | 409 hCoV-19/India/MH-ACTREC-669/2020\|EPI_ISL_700295\|20 | ···················· | ···················· | ···················· | ···················· | ···················· | ···················· |
| W | 410 hCoV-19/India/MH-ACTREC-670/2020\|EPI_ISL_700296\|20 | ···················· | ···················· | ···················· | ···················· | ···················· | ···················· |
| W | 411 hCoV-19/India/MH-ACTREC-671/2020\|EPI_ISL_700297\|20 | ···················· | ···················· | ···················· | ···················· | ···················· | ···················· |
| W | 412 hCoV-19/India/MH-ACTREC-672/2020\|EPI_ISL_700298\|20 | ···················· | ···················· | ···················· | ···················· | ···················· | ···················· |
| W | 413 hCoV-19/India/MH-ACTREC-673/2020\|EPI_ISL_700299\|20 | ···················· | ·A·················· | ···················· | ···················· | ···················· | ··A················· |
| W | 414 hCoV-19/England/ALDP-BB8E3F/2020\|EPI_ISL_702807\|20 | ···················· | ···················· | ···················· | ···················· | ···················· | ···················· |
| W | 415 hCoV-19/England/ALDP-BB942D/2020\|EPI_ISL_702808\|20 | ···················· | ···················· | ···················· | ···················· | ···················· | ···················· |
| W | 416 hCoV-19/England/ALDP-BB54A9/2020\|EPI_ISL_702810\|20 | ···················· | ···················· | ···················· | ···················· | ···················· | ··A················· |
| W | 417 hCoV-19/England/ALDP-BB99E2/2020\|EPI_ISL_702811\|20 | ···················· | ···················· | ···················· | ···················· | ···················· | ···················· |
| W | 418 hCoV-19/England/ALDP-BB2B2F/2020\|EPI_ISL_702813\|20 | ···················· | ·A·················· | ···················· | ···················· | ···················· | ···················· |
| W | 419 hCoV-19/England/ALDP-BB9D6E/2020\|EPI_ISL_702814\|20 | ···················· | ···················· | ···················· | ···················· | ···················· | ···················· |
| W | 420 hCoV-19/England/ALDP-BB57A0/2020\|EPI_ISL_702818\|20 | ···················· | ···················· | ···················· | ···················· | ···················· | ···················· |
| W | 421 hCoV-19/England/ALDP-BB32CF/2020\|EPI_ISL_702821\|20 | ···················· | ···················· | ···················· | ···················· | ···················· | ···················· |
| W | 422 hCoV-19/England/ALDP-BB9A76/2020\|EPI_ISL_702822\|20 | ···················· | ·A·················· | ···················· | ···················· | ···················· | ··A················· |
| W | 423 hCoV-19/England/ALDP-BB9803/2020\|EPI_ISL_702899\|20 | ···················· | ···················· | ···················· | ···················· | ···················· | ···················· |
| W | 424 hCoV-19/England/CAMC-BC0B5B/2020\|EPI_ISL_705058\|20 | ···················· | ···················· | ···················· | ···················· | ···················· | ···················· |
| W | 425 hCoV-19/England/CAMC-BC0827/2020\|EPI_ISL_705080\|20 | ···················· | ···················· | ···················· | ···················· | ···················· | ···················· |
| W | 426 hCoV-19/England/CAMC-BC0C2B/2020\|EPI_ISL_705084\|20 | ···················· | ···················· | ···················· | ···················· | ···················· | ···················· |
| W | 427 hCoV-19/England/CAMC-BC08DC/2020\|EPI_ISL_705085\|20 | ···················· | ···················· | ···················· | ···················· | ···················· | ···················· |
| W | 428 hCoV-19/England/CAMC-BC0159/2020\|EPI_ISL_705087\|20 | ·······T············ | ···················· | ···················· | ···················· | ···················· | ···················· |
| W | 429 hCoV-19/England/CAMC-BC0177/2020\|EPI_ISL_705089\|20 | ·······T············ | ···················· | ···················· | ···················· | ···················· | ···················· |
| W | 430 hCoV-19/England/CAMC-BC112B/2020\|EPI_ISL_705090\|20 | ···················· | ·A·················· | ···················· | ···················· | ···················· | ··A················· |
| W | 431 hCoV-19/England/CAMC-BC1C57/2020\|EPI_ISL_705092\|20 | ···················· | ···················· | ···················· | ···················· | ···················· | ···················· |
| W | 432 hCoV-19/England/CAMC-BC05F3/2020\|EPI_ISL_705093\|20 | ···················· | ·A·················· | ···················· | ···················· | ···················· | ···················· |
| W | 433 hCoV-19/England/CAMC-BC1F21/2020\|EPI_ISL_705095\|20 | ···················· | ···················· | ···················· | ···················· | ···················· | ···················· |
| W | 434 hCoV-19/England/CAMC-BC0E9E/2020\|EPI_ISL_705097\|20 | ···················· | ···················· | ···················· | ···················· | ···················· | ···················· |
| W | 435 hCoV-19/England/CAMC-BC0E52/2020\|EPI_ISL_705098\|20 | ···················· | ···················· | ···················· | ···················· | ···················· | ···················· |
| W | 436 hCoV-19/England/CAMC-BC1ED9/2020\|EPI_ISL_705100\|20 | ···················· | ···················· | ···················· | ···················· | ·····T·············· | ···················· |
| W | 437 hCoV-19/England/CAMC-BC0BB5/2020\|EPI_ISL_705107\|20 | ···················· | ···················· | ···················· | ···················· | ···················· | ···················· |
| W | 438 hCoV-19/England/CAMC-BC01D1/2020\|EPI_ISL_705109\|20 | ···················· | ···················· | ···················· | ···················· | ···················· | ···················· |
| W | 439 hCoV-19/England/CAMC-BBFFCC/2020\|EPI_ISL_705111\|20 | ···················· | ···················· | ···················· | ···················· | ···················· | ···················· |
| W | 440 hCoV-19/England/CAMC-BC2324/2020\|EPI_ISL_705116\|20 | ···················· | ···················· | ···················· | ···················· | ···················· | ···················· |
| W | 441 hCoV-19/England/CAMC-BC0F31/2020\|EPI_ISL_705117\|20 | ···················· | ···················· | ···················· | ···················· | ···················· | ···················· |
| W | 442 hCoV-19/England/NORW-F6F3D/2020\|EPI_ISL_706384\|202 | ···················· | ·A·················· | ···················· | ···················· | ···················· | ··A················· |
| W | 443 hCoV-19/England/NORW-F6F4C/2020\|EPI_ISL_706385\|202 | ···················· | ···················· | ···················· | ···················· | ···················· | ···················· |
| W | 444 hCoV-19/England/NORW-F700C/2020\|EPI_ISL_706388\|202 | ···················· | ···················· | ···················· | ···················· | ···················· | ···················· |
| W | 445 hCoV-19/England/NORW-F7057/2020\|EPI_ISL_706389\|202 | ···················· | ···················· | ···················· | ···················· | ···················· | ···················· |
| W | 446 hCoV-19/England/NORW-F76FB/2020\|EPI_ISL_706400\|202 | ···················· | ···················· | ···················· | ···················· | ···················· | ···················· |
| W | 447 hCoV-19/England/NORW-F7716/2020\|EPI_ISL_706401\|202 | ···················· | ···················· | ···················· | ···················· | ···················· | ···················· |
| W | 448 hCoV-19/England/NORW-F8B82/2020\|EPI_ISL_706402\|202 | ···················· | ···················· | ···················· | ···················· | ···················· | ···················· |
| W | 449 hCoV-19/England/NORW-F8D04/2020\|EPI_ISL_706403\|202 | ···················· | ···················· | ···················· | ···················· | ···················· | ···················· |
| W | 450 hCoV-19/England/NORW-F8D8C/2020\|EPI_ISL_706404\|202 | ···················· | ···················· | ···················· | ···················· | ···················· | ···················· |
| W | 451 hCoV-19/England/NORW-F8F4A/2020\|EPI_ISL_706405\|202 | ···················· | ···················· | ···················· | ···················· | ···················· | ···················· |
| W | 452 hCoV-19/England/NORW-F8F59/2020\|EPI_ISL_706406\|202 | ···················· | ···················· | ···················· | ···················· | ···················· | ···················· |
| W | 453 hCoV-19/England/NORW-F8F68/2020\|EPI_ISL_706407\|202 | ···················· | ···················· | ···················· | ···················· | ···················· | ···················· |
| W | 454 hCoV-19/England/NORW-F8FE0/2020\|EPI_ISL_706408\|202 | ···················· | ···················· | ···················· | ···················· | ···················· | ···················· |
| W | 455 hCoV-19/England/NORW-F9222/2020\|EPI_ISL_706409\|202 | ···················· | ···················· | ···················· | ···················· | ···················· | ···················· |
| W | 456 hCoV-19/England/NOTT-11ECEC/2020\|EPI_ISL_706430\|20 | ···················· | ···················· | ···················· | ···················· | ···················· | ···················· |
| W | 457 hCoV-19/England/NOTT-11F3F5/2020\|EPI_ISL_706431\|20 | ···················· | ···················· | ···················· | ···················· | ···················· | ···················· |
| W | 461 hCoV-19/England/NOTT-1201BB/2020\|EPI_ISL_706435\|20 | ···················· | ···················· | ···················· | ···················· | ···················· | ···················· |
| W | 462 hCoV-19/England/NOTT-1201CA/2020\|EPI_ISL_706436\|20 | ···················· | ···················· | ···················· | ···················· | ···················· | ···················· |
| W | 463 hCoV-19/England/NOTT-1202F4/2020\|EPI_ISL_706437\|20 | ···················· | ···················· | ···················· | ···················· | ···················· | ···················· |
| W | 466 hCoV-19/Wales/PHWC-490010/2020\|EPI_ISL_706440\|2020 | ···················· | ···················· | ···················· | ···················· | ···················· | ···················· |
| W | 469 hCoV-19/Wales/PHWC-490098/2020\|EPI_ISL_706443\|2020 | ···················· | ···················· | ···················· | ···················· | ···················· | ···················· |
| W | 471 hCoV-19/Wales/PHWC-4900F2/2020\|EPI_ISL_706445\|2020 | ···················· | ···················· | ···················· | ···················· | ···················· | ···················· |
| W | 472 hCoV-19/Wales/PHWC-49010E/2020\|EPI_ISL_706446\|2020 | ···················· | ···················· | ···················· | ···················· | ···················· | ···················· |
| W | 474 hCoV-19/Wales/PHWC-49013B/2020\|EPI_ISL_706448\|2020 | ···················· | ···················· | ···················· | ···················· | ···················· | ···················· |
| W | 476 hCoV-19/Tunisia/61600/2020\|EPI_ISL_707700\|2020-11- | ···················· | ···················· | ···················· | ···················· | ···················· | ···················· |
| W | 477 hCoV-19/England/204961132/2020\|EPI_ISL_707703\|2020 | ···················· | ···················· | ···················· | ···················· | ···················· | ···················· |

W 478 hCoV-19/England/204961133/2020|EPI_ISL_707704|2020 .................... .A.................. .................... .................... .................... ..A.................
W 483 hCoV-19/Belgium/ULG-11004/2020|EPI_ISL_707721|2020 .................... .................... .................... .................... .................... ....................
W 484 hCoV-19/Belgium/ULG-10971/2020|EPI_ISL_707723|2020 .................... .................... .................... .................... .................... ....................
W 485 hCoV-19/Belgium/ULG-10983/2020|EPI_ISL_707724|2020 .................... .................... .................... .................... .................... ....................
W 486 hCoV-19/Belgium/ULG-10994/2020|EPI_ISL_707726|2020 .................... .................... .................... .................... .................... ....................
W 487 hCoV-19/Belgium/ULG-10948/2020|EPI_ISL_707727|2020 .................... .................... .................... .................... .................... ....................
W 488 hCoV-19/Belgium/ULG-10949/2020|EPI_ISL_707728|2020 .................... .................... .................... .................... .................... ....................
W 489 hCoV-19/Belgium/ULG-10950/2020|EPI_ISL_707729|2020 .................... .................... .................... .................... .................... ....................
W 490 hCoV-19/Belgium/ULG-10951/2020|EPI_ISL_707730|2020 .................... .................... .................... .................... .................... ....................
W 491 hCoV-19/Belgium/ULG-10952/2020|EPI_ISL_707731|2020 .................... .................... .................... .................... .................... ....................
W 492 hCoV-19/Belgium/ULG-10953/2020|EPI_ISL_707732|2020 .................... .................... .................... .................... .................... ....................
W 493 hCoV-19/Belgium/ULG-10954/2020|EPI_ISL_707733|2020 .................... ...A................ .................... .................... .................... ....................
W 494 hCoV-19/Belgium/ULG-10955/2020|EPI_ISL_707734|2020 .................... .................... .................... .................... .................... ....................
W 495 hCoV-19/Belgium/ULG-10956/2020|EPI_ISL_707735|2020 .................... .................... .................... .................... .................... ....................
W 496 hCoV-19/Belgium/ULG-10957/2020|EPI_ISL_707736|2020 .................... .................... .................... .................... .................... ....................
W 497 hCoV-19/Belgium/ULG-10958/2020|EPI_ISL_707737|2020 .................... .................... .................... .................... .................... ....................
W 498 hCoV-19/Belgium/ULG-10959/2020|EPI_ISL_707738|2020 .................... .................... .................... .................... .................... ....................
W 499 hCoV-19/Belgium/ULG-10960/2020|EPI_ISL_707739|2020 .................... .A.................. .................... .................... .................... ....................
W 500 hCoV-19/Belgium/ULG-10961/2020|EPI_ISL_707740|2020 .................... .................... .................... .................... .................... ....................
W 501 hCoV-19/Belgium/ULG-10965/2020|EPI_ISL_707744|2020 .................... .................... .................... .................... .................... ....................
W 502 hCoV-19/Belgium/ULG-10991/2020|EPI_ISL_707760|2020 .................... .................... .................... .................... .................... ....................
W 503 hCoV-19/Belgium/ULG-10993/2020|EPI_ISL_707761|2020 .................... .................... .................... .................... .................... ....................
W 504 hCoV-19/Belgium/ULG-10995/2020|EPI_ISL_707762|2020 .................... .................... .................... .................... .................... ....................
W 505 hCoV-19/Belgium/ULG-10996/2020|EPI_ISL_707763|2020 .................... .A.................. .................... .................... .................... ..A.................
W 506 hCoV-19/Belgium/ULG-10997/2020|EPI_ISL_707764|2020 .................... .................... .................... .................... .................... ....................
W 507 hCoV-19/Belgium/ULG-10999/2020|EPI_ISL_707765|2020 .................... .................... .................... .................... .................... ....................
W 508 hCoV-19/Belgium/ULG-11000/2020|EPI_ISL_707766|2020 .................... .................... .................... .................... .................... ....................
W 509 hCoV-19/Belgium/ULG-11002/2020|EPI_ISL_707767|2020 .................... .................... .................... .................... .................... ....................
W 510 hCoV-19/Belgium/ULG-11005/2020|EPI_ISL_707768|2020 .................... .................... .................... .................... .................... ....................
W 511 hCoV-19/Belgium/ULG-11010/2020|EPI_ISL_707769|2020 .................... .................... .................... .................... .................... ....................
W 512 hCoV-19/Belgium/ULG-11011/2020|EPI_ISL_707770|2020 .................... .................... .................... .................... .................... ....................
W 513 hCoV-19/USA/AZ-TG445157/2020|EPI_ISL_695000|2020-0 .................... .A.................. .................... .................... .................... ....................
W 514 hCoV-19/USA/AZ-TG445197/2020|EPI_ISL_695002|2020-0 .................... .................... .................... .................... .................... ....................
W 515 hCoV-19/USA/AZ-TG446582/2020|EPI_ISL_695005|2020-0 .................... .................... .................... .................... .................... ....................
W 516 hCoV-19/USA/AZ-TG447447/2020|EPI_ISL_695009|2020-0 .................... .................... .................... .................... .................... ....................
W 517 hCoV-19/USA/AZ-TG447963/2020|EPI_ISL_695013|2020-0 .................... .................... .................... .................... .................... ....................
W 518 hCoV-19/USA/AZ-TG447995/2020|EPI_ISL_695014|2020-0 .................... .A.................. .................... .................... .................... ....................
W 519 hCoV-19/USA/AZ-TG448276/2020|EPI_ISL_695015|2020-0 .................... .................... .................... .................... .................... ....................
W 520 hCoV-19/USA/AZ-TG448488/2020|EPI_ISL_695017|2020-0 .................... .................... .................... .................... .................... ....................
W 521 hCoV-19/USA/AZ-TG448874/2020|EPI_ISL_695019|2020-0 .................... .................... .................... .................... .................... ....................
W 522 hCoV-19/USA/AZ-TG449194/2020|EPI_ISL_695020|2020-0 .................... .................... .................... .................... .................... ....................
W 523 hCoV-19/USA/AZ-TG450430/2020|EPI_ISL_695021|2020-0 .................... .................... .................... .................... .................... ....................
W 524 hCoV-19/USA/AZ-TG452448/2020|EPI_ISL_695022|2020-0 .................... .A.................. .................... .................... .................... ....................
W 525 hCoV-19/USA/AZ-TG452899/2020|EPI_ISL_695025|2020-0 .................... .................... .................... .................... .................... ..N.................
W 526 hCoV-19/USA/AZ-TG452920/2020|EPI_ISL_695026|2020-0 .................... .................... .................... .................... .................... ....................
W 527 hCoV-19/USA/AZ-TG453108/2020|EPI_ISL_695027|2020-0 .................... .................... .................... .................... .................... ....................
W 528 hCoV-19/USA/AZ-TG453802/2020|EPI_ISL_695029|2020-0 .................... .................... .................... .................... .................... ....................
W 529 hCoV-19/USA/AZ-TG454108/2020|EPI_ISL_695031|2020-0 .................... .................... .................... .................... .................... ....................
W 530 hCoV-19/USA/AZ-TG454541/2020|EPI_ISL_695034|2020-0 .................... .A.................. .................... .................... .................... ....................
W 531 hCoV-19/USA/AZ-TG455129/2020|EPI_ISL_695035|2020-0 .................... .................... .................... .................... .................... ....................
W 532 hCoV-19/USA/AZ-TG455598/2020|EPI_ISL_695036|2020-0 .................... .................... .................... .................... .................... ....................
W 533 hCoV-19/USA/AZ-TG455599/2020|EPI_ISL_695037|2020-0 .................... .A.................. .................... .................... .................... ....................
W 534 hCoV-19/USA/AZ-TG455737/2020|EPI_ISL_695039|2020-0 .................... .................... .................... .................... .................... ....................
W 535 hCoV-19/USA/AZ-TG456066/2020|EPI_ISL_695041|2020-0 .................... .................... .................... .................... .................... ....................
W 536 hCoV-19/USA/AZ-TG456323/2020|EPI_ISL_695042|2020-0 .................... .A.................. .................... .................... .................... ....................
W 537 hCoV-19/USA/AZ-TG456402/2020|EPI_ISL_695043|2020-0 .................... .A.................. .................... .................... .................... ....................
W 538 hCoV-19/USA/AZ-TG458924/2020|EPI_ISL_695047|2020-0 .................... .A.................. .................... .................... .................... ....................
W 539 hCoV-19/USA/AZ-TG670109/2020|EPI_ISL_696327|2020-0 .................... .................... .................... .................... .................... ....................
W 540 hCoV-19/USA/AZ-TG670110/2020|EPI_ISL_696328|2020-0 .................... .................... .................... .................... .................... ....................
W 541 hCoV-19/USA/AZ-TG670119/2020|EPI_ISL_696335|2020-0 .................... .................... .................... .................... .................... ....................
W 542 hCoV-19/USA/AZ-TG670125/2020|EPI_ISL_696338|2020-0 .................... .................... .................... .................... .................... ....................
W 543 hCoV-19/USA/AZ-TG670131/2020|EPI_ISL_696342|2020-0 .................... .................... .................... .................... .................... ....................
W 544 hCoV-19/USA/AZ-TG670136/2020|EPI_ISL_696345|2020-0 .................... .................... .................... .................... .................... ....................
W 545 hCoV-19/USA/AZ-TG670141/2020|EPI_ISL_696348|2020-0 .................... .................... .................... .................... .................... ....................
W 546 hCoV-19/USA/AZ-TG670151/2020|EPI_ISL_696354|2020-0 .................... .................... .................... .................... .................... ....................
W 547 hCoV-19/USA/AZ-TG670168/2020|EPI_ISL_696366|2020-0 .................... .................... .................... .................... .................... ....................
W 548 hCoV-19/USA/AZ-TG670171/2020|EPI_ISL_696369|2020-0 .................... .................... .................... .................... .................... ....................
W 549 hCoV-19/USA/AZ-TG670177/2020|EPI_ISL_696374|2020-0 .................... .................... .................... .................... .................... ....................
W 550 hCoV-19/USA/AZ-TG670178/2020|EPI_ISL_696375|2020-0 .................... .................... .................... .................... .................... ....................
W 551 hCoV-19/USA/AZ-TG670179/2020|EPI_ISL_696376|2020-0 .................... .................... .................... .................... .................... ....................
W 552 hCoV-19/USA/AZ-TG670181/2020|EPI_ISL_696377|2020-0 .................... .................... .................... .................... .................... ....................
W 553 hCoV-19/United Arab Emirates/0751/2020|EPI_ISL_698 .................... .................... .................... .................... .................... ....................
W 554 hCoV-19/United Arab Emirates/0752/2020|EPI_ISL_698 .................... .................... .................... .................... .................... ....................
W 555 hCoV-19/United Arab Emirates/0754/2020|EPI_ISL_698 .................... .................... .................... .................... .................... ....................
W 556 hCoV-19/United Arab Emirates/0755/2020|EPI_ISL_698 .................... .................... .................... .................... .................... ....................
W 557 hCoV-19/United Arab Emirates/0756/2020|EPI_ISL_698 .................... .A.................. .................... .................... .................... ....................
W 558 hCoV-19/United Arab Emirates/0759/2020|EPI_ISL_698 .................... .................... .................... .................... .................... ....................
W 559 hCoV-19/United Arab Emirates/0760/2020|EPI_ISL_698 .................... .................... .................... .................... .................... ....................
W 560 hCoV-19/United Arab Emirates/0762/2020|EPI_ISL_698 .................... .................... .................... .................... .................... ....................
W 561 hCoV-19/United Arab Emirates/0764/2020|EPI_ISL_698 .................... .................... .................... .................... .................... ....................

| | | | | | | | | |
|---|---|---|---|---|---|---|---|---|
| W | 562 | hCoV-19/United Arab Emirates/0765/2020\|EPI_ISL_698 | ···················· | ···················· | ····················· | ···················· | ···················· | ···················· |
| W | 563 | hCoV-19/United Arab Emirates/0766/2020\|EPI_ISL_698 | ···················· | ···················· | ····················· | ···················· | ···················· | ···················· |
| W | 564 | hCoV-19/United Arab Emirates/0767/2020\|EPI_ISL_698 | ···················· | ···················· | ····················· | ···················· | ···················· | ···················· |
| W | 565 | hCoV-19/United Arab Emirates/0768/2020\|EPI_ISL_698 | ···················· | ···················· | ····················· | ···················· | ···················· | ···················· |
| W | 566 | hCoV-19/United Arab Emirates/0769/2020\|EPI_ISL_698 | ···················· | ···················· | ····················· | ···················· | ···················· | ···················· |
| W | 567 | hCoV-19/United Arab Emirates/0771/2020\|EPI_ISL_698 | ···················· | ···················· | ····················· | ···················· | ···················· | ···················· |
| W | 568 | hCoV-19/United Arab Emirates/0772/2020\|EPI_ISL_698 | ···················· | ···················· | ····················· | ···················· | ···················· | ···················· |
| W | 569 | hCoV-19/United Arab Emirates/0773/2020\|EPI_ISL_698 | ···················· | ···················· | ····················· | ···················· | ···················· | ···················· |
| W | 570 | hCoV-19/United Arab Emirates/0774/2020\|EPI_ISL_698 | ···················· | ···················· | ····················· | ···················· | ···················· | ···················· |
| W | 571 | hCoV-19/United Arab Emirates/0775/2020\|EPI_ISL_698 | ···················· | ···················· | ····················· | ···················· | ···················· | ···················· |
| W | 572 | hCoV-19/United Arab Emirates/0776/2020\|EPI_ISL_698 | ···················· | ···················· | ····················· | ···················· | ···················· | ···················· |
| W | 573 | hCoV-19/United Arab Emirates/0777/2020\|EPI_ISL_698 | ···················· | ···················· | ····················· | ···················· | ···················· | ···················· |
| W | 574 | hCoV-19/United Arab Emirates/0778/2020\|EPI_ISL_698 | ···················· | ···················· | ····················· | ···················· | ···················· | ···················· |
| W | 575 | hCoV-19/United Arab Emirates/0779/2020\|EPI_ISL_698 | ···················· | ···················· | ····················· | ···················· | ···················· | ···················· |
| W | 576 | hCoV-19/United Arab Emirates/0780/2020\|EPI_ISL_698 | ···················· | ···················· | ····················· | ···················· | ···················· | ···················· |
| W | 577 | hCoV-19/United Arab Emirates/0781/2020\|EPI_ISL_698 | ···················· | ···················· | ····················· | ···················· | ···················· | ···················· |
| W | 578 | hCoV-19/United Arab Emirates/0783/2020\|EPI_ISL_698 | ···················· | ···················· | ····················· | ···················· | ···················· | ···················· |
| W | 579 | hCoV-19/United Arab Emirates/0784/2020\|EPI_ISL_698 | ···················· | ···················· | ····················· | ···················· | ···················· | ···················· |
| W | 580 | hCoV-19/United Arab Emirates/0786/2020\|EPI_ISL_698 | ···················· | ···················· | ····················· | ···················· | ···················· | ···················· |
| W | 581 | hCoV-19/United Arab Emirates/0787/2020\|EPI_ISL_698 | ···················· | ···················· | ····················· | ···················· | ···················· | ···················· |
| W | 582 | hCoV-19/United Arab Emirates/0789/2020\|EPI_ISL_698 | ···················· | ···················· | ····················· | ···················· | ···················· | ···················· |
| W | 583 | hCoV-19/United Arab Emirates/0790/2020\|EPI_ISL_698 | ···················· | ···················· | ····················· | ···················· | ···················· | ···················· |
| W | 584 | hCoV-19/United Arab Emirates/0791/2020\|EPI_ISL_698 | ···················· | ···················· | ····················· | ···················· | ···················· | ···················· |
| W | 585 | hCoV-19/United Arab Emirates/0792/2020\|EPI_ISL_698 | ···················· | ···················· | ····················· | ···················· | ···················· | ···················· |
| W | 586 | hCoV-19/United Arab Emirates/0793/2020\|EPI_ISL_698 | ········T··········· | ···················· | ····················· | ···················· | ···················· | ···················· |
| W | 587 | hCoV-19/United Arab Emirates/0794/2020\|EPI_ISL_698 | ···················· | ···················· | ····················· | ···················· | ···················· | ···················· |
| W | 588 | hCoV-19/United Arab Emirates/0795/2020\|EPI_ISL_698 | ···················· | ···················· | ····················· | ···················· | ···················· | ···················· |
| W | 589 | hCoV-19/United Arab Emirates/0796/2020\|EPI_ISL_698 | ···················· | ···················· | ····················· | ···················· | ···················· | ···················· |
| W | 590 | hCoV-19/United Arab Emirates/0797/2020\|EPI_ISL_698 | ···················· | ···················· | ····················· | ···················· | ···················· | ···················· |
| W | 591 | hCoV-19/United Arab Emirates/0798/2020\|EPI_ISL_698 | ···················· | ···················· | ····················· | ···················· | ···················· | ···················· |
| W | 592 | hCoV-19/United Arab Emirates/0799/2020\|EPI_ISL_698 | ···················· | ···················· | ····················· | ···················· | ···················· | ···················· |
| W | 593 | hCoV-19/United Arab Emirates/0800/2020\|EPI_ISL_698 | ···················· | ···················· | ····················· | ···················· | ···················· | ···················· |
| W | 594 | hCoV-19/United Arab Emirates/0801/2020\|EPI_ISL_698 | ···················· | ···················· | ····················· | ···················· | ···················· | ···················· |
| W | 595 | hCoV-19/United Arab Emirates/0841/2020\|EPI_ISL_698 | ···················· | ···················· | ····················· | ···················· | ···················· | ···················· |
| W | 596 | hCoV-19/United Arab Emirates/0842/2020\|EPI_ISL_698 | ···················· | ···················· | ····················· | ···················· | ···················· | ···················· |
| W | 597 | hCoV-19/United Arab Emirates/0847/2020\|EPI_ISL_698 | ···················· | ···················· | ····················· | ···················· | ···················· | ···················· |
| W | 598 | hCoV-19/United Arab Emirates/0848/2020\|EPI_ISL_698 | ···················· | ···················· | ····················· | ···················· | ···················· | ···················· |
| W | 599 | hCoV-19/United Arab Emirates/0849/2020\|EPI_ISL_698 | ···················· | ···················· | ····················· | ···················· | ···················· | ···················· |
| W | 600 | hCoV-19/United Arab Emirates/0850/2020\|EPI_ISL_698 | ···················· | ···················· | ····················· | ···················· | ···················· | ···················· |
| W | 601 | hCoV-19/United Arab Emirates/0851/2020\|EPI_ISL_698 | ···················· | ···················· | ····················· | ···················· | ···················· | ···················· |
| W | 602 | hCoV-19/United Arab Emirates/0852/2020\|EPI_ISL_698 | ···················· | ···················· | ····················· | ···················· | ···················· | ···················· |
| W | 603 | hCoV-19/United Arab Emirates/0853/2020\|EPI_ISL_698 | ···················· | ···················· | ····················· | ···················· | ···················· | ···················· |
| W | 604 | hCoV-19/United Arab Emirates/0854/2020\|EPI_ISL_698 | ···················· | ···················· | ····················· | ···················· | ···················· | ···················· |
| W | 605 | hCoV-19/United Arab Emirates/0855/2020\|EPI_ISL_698 | ···················· | ···················· | ····················· | ···················· | ···················· | ···················· |
| W | 606 | hCoV-19/United Arab Emirates/0856/2020\|EPI_ISL_698 | ···················· | ···················· | ····················· | ···················· | ···················· | ···················· |
| W | 607 | hCoV-19/United Arab Emirates/0857/2020\|EPI_ISL_698 | ···················· | ···················· | ····················· | ···················· | ···················· | ···················· |
| W | 608 | hCoV-19/United Arab Emirates/0858/2020\|EPI_ISL_698 | ···················· | ···················· | ····················· | ···················· | ···················· | ···················· |
| W | 609 | hCoV-19/United Arab Emirates/0859/2020\|EPI_ISL_698 | ···················· | ···················· | ····················· | ···················· | ···················· | ···················· |
| W | 610 | hCoV-19/United Arab Emirates/0860/2020\|EPI_ISL_698 | ···················· | ···················· | ····················· | ···················· | ···················· | ···················· |
| W | 611 | hCoV-19/India/MH-ACTREC-675/2020\|EPI_ISL_700300\|20 | ···················· | ···················· | ····················· | ···················· | ···················· | ···················· |
| W | 612 | hCoV-19/India/MH-ACTREC-676/2020\|EPI_ISL_700301\|20 | ···················· | ···················· | ····················· | ···················· | ···················· | ···················· |
| W | 613 | hCoV-19/India/MH-ACTREC-677/2020\|EPI_ISL_700302\|20 | ···················· | ···················· | ····················· | ···················· | ···················· | ···················· |
| W | 614 | hCoV-19/India/MH-ACTREC-678/2020\|EPI_ISL_700303\|20 | ···················· | ···················· | ····················· | ···················· | ···················· | ···················· |
| W | 615 | hCoV-19/India/MH-ACTREC-679/2020\|EPI_ISL_700304\|20 | ···················· | ···················· | ····················· | ···················· | ···················· | ···················· |
| W | 616 | hCoV-19/India/MH-ACTREC-680/2020\|EPI_ISL_700305\|20 | ···················· | ···················· | ····················· | ···················· | ···················· | ···················· |
| W | 617 | hCoV-19/England/ALDP-BB98E5/2020\|EPI_ISL_702901\|20 | ···················· | ···················· | ····················· | ···················· | ···················· | ···················· |
| W | 618 | hCoV-19/England/ALDP-BB9A67/2020\|EPI_ISL_702903\|20 | ···················· | ···················· | ····················· | ···················· | ···················· | ···················· |
| W | 619 | hCoV-19/England/ALDP-BB2A6E/2020\|EPI_ISL_702906\|20 | ···················· | ···················· | ····················· | ···················· | ···················· | ···················· |
| W | 620 | hCoV-19/England/ALDP-BB9CCB/2020\|EPI_ISL_702909\|20 | ···················· | ···················· | ····················· | ···················· | ···················· | ···················· |
| W | 621 | hCoV-19/England/ALDP-BB9BFB/2020\|EPI_ISL_702912\|20 | ···················· | ···················· | ····················· | ···················· | ···················· | ···················· |
| W | 622 | hCoV-19/England/ALDP-BB599B/2020\|EPI_ISL_702914\|20 | ···················· | ···················· | ····················· | ···················· | ···················· | ···················· |
| W | 623 | hCoV-19/England/ALDP-BB39E8/2020\|EPI_ISL_702916\|20 | ···················· | ···················· | ····················· | ···················· | ······T············· | ···················· |
| W | 624 | hCoV-19/England/ALDP-BB52FA/2020\|EPI_ISL_702919\|20 | ···················· | ···················· | ····················· | ···················· | ···················· | ···················· |
| W | 625 | hCoV-19/England/ALDP-BB988B/2020\|EPI_ISL_702921\|20 | ···················· | ···················· | ····················· | ···················· | ···················· | ···················· |
| W | 626 | hCoV-19/England/ALDP-BB3353/2020\|EPI_ISL_702923\|20 | ···················· | ···················· | ····················· | ···················· | ······T············· | ···················· |
| W | 627 | hCoV-19/England/ALDP-BB9CBC/2020\|EPI_ISL_702924\|20 | ···················· | ···················· | ····················· | ···················· | ···················· | ···················· |
| W | 628 | hCoV-19/England/ALDP-BB28CE/2020\|EPI_ISL_702929\|20 | ···················· | ···················· | ····················· | ···················· | ···················· | ···················· |
| W | 629 | hCoV-19/England/ALDP-BB59B9/2020\|EPI_ISL_702931\|20 | ···················· | ···················· | ····················· | ···················· | ···················· | ···················· |
| W | 630 | hCoV-19/England/ALDP-BB348D/2020\|EPI_ISL_702933\|20 | ···················· | ···················· | ····················· | ···················· | ···················· | ···················· |
| W | 631 | hCoV-19/England/ALDP-BB5597/2020\|EPI_ISL_702936\|20 | ···················· | ···················· | ····················· | ···················· | ···················· | ···················· |
| W | 632 | hCoV-19/England/ALDP-BB27EF/2020\|EPI_ISL_702938\|20 | ···················· | ···················· | ····················· | ···················· | ···················· | ···················· |
| W | 633 | hCoV-19/England/ALDP-BB556A/2020\|EPI_ISL_702948\|20 | ···················· | ···················· | ····················· | ···················· | ···················· | ···················· |
| W | 634 | hCoV-19/England/ALDP-BB2864/2020\|EPI_ISL_702951\|20 | ···················· | ···················· | ····················· | ···················· | ···················· | ···················· |
| W | 635 | hCoV-19/England/ALDP-BB2E08/2020\|EPI_ISL_702954\|20 | ···················· | ···················· | ····················· | ···················· | ······T············· | ···················· |
| W | 636 | hCoV-19/England/ALDP-BB5746/2020\|EPI_ISL_702955\|20 | ···················· | ···················· | ····················· | ···················· | ···················· | ···················· |
| W | 637 | hCoV-19/England/ALDP-BB9CF8/2020\|EPI_ISL_702957\|20 | ···················· | ···················· | ····················· | ···················· | ···················· | ···················· |
| W | 638 | hCoV-19/England/ALDP-BB995B/2020\|EPI_ISL_702958\|20 | ···················· | ···················· | ····················· | ···················· | ···················· | ···················· |
| W | 639 | hCoV-19/England/ALDP-BB9B64/2020\|EPI_ISL_702960\|20 | ···················· | ·A·················· | ····················· | ···················· | ···················· | ···················· |
| W | 640 | hCoV-19/England/ALDP-BB2A5F/2020\|EPI_ISL_702967\|20 | ···················· | ···················· | ····················· | ···················· | ······T············· | ···················· |
| W | 641 | hCoV-19/England/CAMC-BC0098/2020\|EPI_ISL_705122\|20 | ···················· | ···················· | ····················· | ···················· | ···················· | ···················· |

| | | | | | | | | | |
|---|---|---|---|---|---|---|---|---|---|
| W | 642 | hCoV-19/England/CAMC-BC2096/2020\|EPI_ISL_705123\|20 | ···················· | ···················· | ····················· | ···················· | ···················· | ···················· | |
| W | 643 | hCoV-19/England/CAMC-BC1C75/2020\|EPI_ISL_705127\|20 | ···················· | ···················· | ····················· | ···················· | ···················· | ···················· | |
| W | 644 | hCoV-19/England/CAMC-BC0863/2020\|EPI_ISL_705128\|20 | ···················· | ·A·················· | ····················· | ···················· | ···················· | ···················· | |
| W | 645 | hCoV-19/England/CAMC-BC0EE9/2020\|EPI_ISL_705131\|20 | ···················· | ···················· | ····················· | ···················· | ···················· | ···················· | |
| W | 646 | hCoV-19/England/CAMC-BC004D/2020\|EPI_ISL_705135\|20 | ···················· | ···················· | ····················· | ···················· | ···················· | ···················· | |
| W | 647 | hCoV-19/England/CAMC-BC0775/2020\|EPI_ISL_705136\|20 | ···················· | ···················· | ····················· | ···················· | ···················· | ···················· | |
| W | 648 | hCoV-19/England/CAMC-BBFEDE/2020\|EPI_ISL_705141\|20 | ···················· | ·A·················· | ····················· | ···················· | ···················· | ···················· | |
| W | 649 | hCoV-19/England/CAMC-BC0793/2020\|EPI_ISL_705142\|20 | ···················· | ···················· | ····················· | ···················· | ···················· | ···················· | |
| W | 650 | hCoV-19/England/CAMC-BC01FF/2020\|EPI_ISL_705144\|20 | ···················· | ···················· | ····················· | ···················· | ···················· | ···················· | |
| W | 651 | hCoV-19/England/CAMC-BC1F4F/2020\|EPI_ISL_705146\|20 | ···················· | ···················· | ····················· | ···················· | ···················· | ···A················ | |
| W | 652 | hCoV-19/England/CAMC-BC0809/2020\|EPI_ISL_705147\|20 | ········T··········· | ···················· | ····················· | ···················· | ···················· | ···················· | |
| W | 653 | hCoV-19/England/CAMC-BC0F13/2020\|EPI_ISL_705152\|20 | ···················· | ···················· | ····················· | ···················· | ···················· | ···················· | |
| W | 654 | hCoV-19/England/CAMC-BC0678/2020\|EPI_ISL_705159\|20 | ···················· | ···················· | ····················· | ···················· | ···················· | ···················· | |
| W | 655 | hCoV-19/England/CAMC-BC061E/2020\|EPI_ISL_705162\|20 | ···················· | ···················· | ····················· | ···················· | ···················· | ···················· | |
| W | 656 | hCoV-19/England/CAMC-BC0BA6/2020\|EPI_ISL_705165\|20 | ···················· | ···················· | ····················· | ···················· | ······T············· | ···················· | |
| W | 657 | hCoV-19/England/CAMC-BC03CC/2020\|EPI_ISL_705167\|20 | ···················· | ···················· | ····················· | ···················· | ···················· | ···················· | |
| W | 658 | hCoV-19/England/CAMC-BC00B6/2020\|EPI_ISL_705169\|20 | ···················· | ···················· | ····················· | ···················· | ···················· | ···················· | |
| W | 659 | hCoV-19/England/CAMC-BC0F40/2020\|EPI_ISL_705172\|20 | ···················· | ···················· | ····················· | ···················· | ···················· | ···················· | |
| W | 660 | hCoV-19/England/CAMC-BC0766/2020\|EPI_ISL_705175\|20 | ···················· | ···················· | ····················· | ···················· | ···················· | ···················· | |
| W | 661 | hCoV-19/England/CAMC-BBFF08/2020\|EPI_ISL_705177\|20 | ···················· | ···················· | ····················· | ···················· | ···················· | ···················· | |
| W | 662 | hCoV-19/England/CAMC-BC08CD/2020\|EPI_ISL_705180\|20 | ···················· | ···················· | ····················· | ···················· | ···················· | ···················· | |
| W | 663 | hCoV-19/England/CAMC-BC02ED/2020\|EPI_ISL_705181\|20 | ···················· | ···················· | ····················· | ···················· | ···················· | ···················· | |
| W | 664 | hCoV-19/England/CAMC-BC20A5/2020\|EPI_ISL_705183\|20 | ···················· | ···················· | ····················· | ···················· | ···················· | ···················· | |
| W | 665 | hCoV-19/England/CAMC-BC1C66/2020\|EPI_ISL_705185\|20 | ···················· | ···················· | ····················· | ···················· | ···················· | ···················· | |
| W | 666 | hCoV-19/England/CAMC-BC056C/2020\|EPI_ISL_705186\|20 | ···················· | ······A············· | ····················· | ···················· | ···················· | ···················· | |
| W | 667 | hCoV-19/England/CAMC-BC1C1B/2020\|EPI_ISL_705188\|20 | ···················· | ···················· | ····················· | ···················· | ···················· | ···················· | |
| W | 668 | hCoV-19/England/MILK-BAB215/2020\|EPI_ISL_705191\|20 | ···················· | ···················· | ····················· | ···················· | ···················· | ···················· | |
| W | 669 | hCoV-19/England/MILK-BA998B/2020\|EPI_ISL_705192\|20 | ···················· | ·A·················· | ····················· | ···················· | ···················· | ···A················ | |
| W | 670 | hCoV-19/England/MILK-BA996D/2020\|EPI_ISL_705196\|20 | ···················· | ···················· | ····················· | ···················· | ···················· | ···················· | |
| W | 671 | hCoV-19/England/MILK-BA9B58/2020\|EPI_ISL_705199\|20 | ···················· | ···················· | ····················· | ···················· | ···················· | ···················· | |
| W | 672 | hCoV-19/England/NORW-F9240/2020\|EPI_ISL_706410\|202 | ···················· | ···················· | ····················· | ···················· | ···················· | ···················· | |
| W | 673 | hCoV-19/England/NORW-F937A/2020\|EPI_ISL_706411\|202 | ···················· | ···················· | ····················· | ···················· | ···················· | ···················· | |
| W | 674 | hCoV-19/England/NORW-F941D/2020\|EPI_ISL_706413\|202 | ···················· | ···················· | ····················· | ···················· | ···················· | ···················· | |
| W | 675 | hCoV-19/England/NORW-F94FF/2020\|EPI_ISL_706414\|202 | ···················· | ···················· | ····················· | ···················· | ···················· | ···················· | |
| W | 676 | hCoV-19/England/NORW-F9538/2020\|EPI_ISL_706415\|202 | ···················· | ···················· | ····················· | ···················· | ···················· | ···················· | |
| W | 677 | hCoV-19/England/NORW-F9626/2020\|EPI_ISL_706416\|202 | ···················· | ···················· | ····················· | ···················· | ···················· | ···················· | |
| W | 678 | hCoV-19/England/NORW-F969F/2020\|EPI_ISL_706417\|202 | ···················· | ···················· | ····················· | ···················· | ···················· | ···················· | |
| W | 679 | hCoV-19/England/NORW-F96EA/2020\|EPI_ISL_706418\|202 | ···················· | ···················· | ····················· | ···················· | ···················· | ···················· | |
| W | 680 | hCoV-19/England/NORW-F9741/2020\|EPI_ISL_706419\|202 | ···················· | ···················· | ····················· | ···················· | ···················· | ···················· | |
| W | 681 | hCoV-19/England/NOTT-11D2DC/2020\|EPI_ISL_706420\|20 | ···················· | ···················· | ····················· | ···················· | ···················· | ···················· | |
| W | 682 | hCoV-19/England/NOTT-11D7BF/2020\|EPI_ISL_706421\|20 | ···················· | ···················· | ····················· | ···················· | ···················· | ···················· | |
| W | 683 | hCoV-19/England/NOTT-11DAF2/2020\|EPI_ISL_706422\|20 | ···················· | ···················· | ····················· | ···················· | ···················· | ···················· | |
| W | 684 | hCoV-19/England/NOTT-11DB0E/2020\|EPI_ISL_706423\|20 | ···················· | ···················· | ····················· | ···················· | ···················· | ···················· | |
| W | 685 | hCoV-19/England/NOTT-11DB3B/2020\|EPI_ISL_706424\|20 | ···················· | ···················· | ····················· | ···················· | ···················· | ···················· | |
| W | 686 | hCoV-19/England/NOTT-11DBFF/2020\|EPI_ISL_706425\|20 | ···················· | ···················· | ····················· | ···················· | ···················· | ···················· | |
| W | 687 | hCoV-19/England/NOTT-11DF7B/2020\|EPI_ISL_706426\|20 | ···················· | ···················· | ····················· | ···················· | ···················· | ···················· | |
| W | 688 | hCoV-19/England/NOTT-11E2AE/2020\|EPI_ISL_706427\|20 | ···················· | ···················· | ····················· | ···················· | ···················· | ···················· | |
| W | 689 | hCoV-19/England/NOTT-11E6A2/2020\|EPI_ISL_706428\|20 | ···················· | ···················· | ····················· | ···················· | ···················· | ···················· | |
| W | 690 | hCoV-19/England/NOTT-11ECCE/2020\|EPI_ISL_706429\|20 | ···················· | ···················· | ····················· | ···················· | ···················· | ···················· | |
| W | 691 | hCoV-19/Wales/PHWC-490168/2020\|EPI_ISL_706450\|2020 | ···················· | ···················· | ····················· | ···················· | ···················· | ···················· | |
| W | 692 | hCoV-19/Wales/PHWC-490186/2020\|EPI_ISL_706451\|2020 | ···················· | ···················· | ····················· | ···················· | ···················· | ···················· | |
| W | 693 | hCoV-19/Wales/PHWC-490195/2020\|EPI_ISL_706452\|2020 | ···················· | ···················· | ····················· | ···················· | ···················· | ···················· | |
| W | 694 | hCoV-19/Wales/PHWC-4901A4/2020\|EPI_ISL_706453\|2020 | ···················· | ···················· | ····················· | ···················· | ···················· | ···················· | |
| W | 695 | hCoV-19/Wales/PHWC-4901B3/2020\|EPI_ISL_706454\|2020 | ···················· | ···················· | ····················· | ···················· | ···················· | ···················· | |
| W | 697 | hCoV-19/Wales/PHWC-4901D1/2020\|EPI_ISL_706456\|2020 | ···················· | ···················· | ····················· | ···················· | ···················· | ···················· | |
| W | 698 | hCoV-19/Wales/PHWC-4901E0/2020\|EPI_ISL_706457\|2020 | ···················· | ···················· | ····················· | ···················· | ···················· | ···················· | |
| W | 699 | hCoV-19/Wales/PHWC-4901FF/2020\|EPI_ISL_706458\|2020 | ···················· | ···················· | ····················· | ···················· | ···················· | ···················· | |
| W | 700 | hCoV-19/Wales/PHWC-49020B/2020\|EPI_ISL_706459\|2020 | ···················· | ···················· | ····················· | ···················· | ····T··············· | ···················· | |
| W | 701 | hCoV-19/Wales/PHWC-49021A/2020\|EPI_ISL_706460\|2020 | ···················· | ···················· | ····················· | ···················· | ····T··············· | ···················· | |
| W | 702 | hCoV-19/Wales/PHWC-490247/2020\|EPI_ISL_706461\|2020 | ···················· | ···················· | ····················· | ···················· | ····T··············· | ···················· | |
| W | 703 | hCoV-19/Wales/PHWC-490256/2020\|EPI_ISL_706462\|2020 | ···················· | ···················· | ····················· | ···················· | ···················· | ···················· | |
| W | 714 | hCoV-19/Wales/PHWC-49039F/2020\|EPI_ISL_706473\|2020 | ···················· | ···················· | ····················· | ···················· | ···················· | ···················· | |
| W | 723 | hCoV-19/Wales/PHWC-49049C/2020\|EPI_ISL_706482\|2020 | ···················· | ···················· | ····················· | ···················· | ···················· | ···················· | |
| W | 726 | hCoV-19/Wales/PHWC-490511/2020\|EPI_ISL_706485\|2020 | ···················· | ···················· | ····················· | ···················· | ···················· | ···················· | |
| W | 728 | hCoV-19/Wales/PHWC-49054E/2020\|EPI_ISL_706487\|2020 | ···················· | ···················· | ····················· | ···················· | ···················· | ···················· | |
| W | 730 | hCoV-19/Wales/PHWC-490599/2020\|EPI_ISL_706489\|2020 | ···················· | ···················· | ····················· | ···················· | ···················· | ···················· | |
| W | 732 | hCoV-19/Wales/PHWC-4905C6/2020\|EPI_ISL_706491\|2020 | ···················· | ···················· | ····················· | ···················· | ···················· | ···················· | |
| W | 733 | hCoV-19/Wales/PHWC-4905E4/2020\|EPI_ISL_706492\|2020 | ···················· | ···················· | ····················· | ···················· | NNN················· | ···················· | 1 |
| W | 736 | hCoV-19/Wales/PHWC-49062D/2020\|EPI_ISL_706495\|2020 | ···················· | ···················· | ····················· | ···················· | ···················· | ···················· | |
| W | 740 | hCoV-19/Wales/PHWC-490687/2020\|EPI_ISL_706499\|2020 | ···················· | ···················· | ····················· | ···················· | ···················· | ···················· | |
| W | 741 | hCoV-19/Belgium/ULG-10962/2020\|EPI_ISL_707741\|2020 | ···················· | ···················· | ····················· | ···················· | ···················· | ···················· | |
| W | 742 | hCoV-19/Belgium/ULG-10963/2020\|EPI_ISL_707742\|2020 | ···················· | ···················· | ····················· | ···················· | ···················· | ···················· | |
| W | 743 | hCoV-19/Belgium/ULG-10964/2020\|EPI_ISL_707743\|2020 | ···················· | ···················· | ····················· | ···················· | ···················· | ···················· | |
| W | 744 | hCoV-19/Belgium/ULG-10966/2020\|EPI_ISL_707745\|2020 | ···················· | ···················· | ····················· | ···················· | ···················· | ···················· | |
| W | 745 | hCoV-19/Belgium/ULG-10968/2020\|EPI_ISL_707746\|2020 | ···················· | ···················· | ····················· | ···················· | ···················· | ···················· | |
| W | 746 | hCoV-19/Belgium/ULG-10970/2020\|EPI_ISL_707747\|2020 | ···················· | ···················· | ····················· | ···················· | ···················· | ···················· | |
| W | 747 | hCoV-19/Belgium/ULG-10972/2020\|EPI_ISL_707748\|2020 | ···················· | ···················· | ····················· | ···················· | ···················· | ···················· | |
| W | 748 | hCoV-19/Belgium/ULG-10973/2020\|EPI_ISL_707749\|2020 | ···················· | ···················· | ····················· | ···················· | ···················· | ···················· | |
| W | 749 | hCoV-19/Belgium/ULG-10975/2020\|EPI_ISL_707750\|2020 | ···················· | ···················· | ····················· | ···················· | ···················· | ···················· | |
| W | 750 | hCoV-19/Belgium/ULG-10977/2020\|EPI_ISL_707751\|2020 | ···················· | ···················· | ····················· | ···················· | ···················· | ···················· | |

| | | | | | | | | |
|---|---|---|---|---|---|---|---|---|
| W | 751 | hCoV-19/Belgium/ULG-10978/2020\|EPI_ISL_707752\|2020 | ···················· | ···················· | ···················· | ···················· | ···················· | ···················· |
| W | 752 | hCoV-19/Belgium/ULG-10979/2020\|EPI_ISL_707753\|2020 | ···················· | ···················· | ···················· | ···················· | ···················· | ···················· |
| W | 753 | hCoV-19/Belgium/ULG-10980/2020\|EPI_ISL_707754\|2020 | ···················· | ·A·················· | ···················· | ···················· | ···················· | ···················· |
| W | 754 | hCoV-19/Belgium/ULG-10984/2020\|EPI_ISL_707755\|2020 | ···················· | ···················· | ···················· | ···················· | ···················· | ···················· |
| W | 755 | hCoV-19/Belgium/ULG-10985/2020\|EPI_ISL_707756\|2020 | ···················· | ···················· | ···················· | ···················· | ···················· | ···················· |
| W | 756 | hCoV-19/Belgium/ULG-10988/2020\|EPI_ISL_707757\|2020 | ···················· | ···················· | ···················· | ···················· | ···················· | ···················· |
| W | 757 | hCoV-19/Belgium/ULG-10989/2020\|EPI_ISL_707758\|2020 | ···················· | ···················· | ···················· | ···················· | ···················· | ···················· |
| W | 758 | hCoV-19/Belgium/ULG-10990/2020\|EPI_ISL_707759\|2020 | ···················· | ···················· | ···················· | ···················· | ···················· | ···················· |
| W | 759 | hCoV-19/France/NAQ-isl1/2020\|EPI_ISL_707781\|2020-0 | ···················· | ···················· | ···················· | ···················· | ···················· | ···················· |
| W | 760 | hCoV-19/France/NAQ-isl2/2020\|EPI_ISL_707782\|2020-0 | ···················· | ···················· | ···················· | ···················· | ···················· | ···················· |
| W | 761 | hCoV-19/Tunisia/2909/2020\|EPI_ISL_707791\|2020-03-0 | ···················· | ···················· | ···················· | ···················· | ···················· | ···················· |
| W | 762 | hCoV-19/Tunisia/6736/2020\|EPI_ISL_707792\|2020-03-3 | ···················· | ···················· | ···················· | ···················· | ···················· | ···················· |
| W | 763 | hCoV-19/Tunisia/9111/2020\|EPI_ISL_707793\|2020-04-1 | ···················· | ···················· | ···················· | ···················· | ···················· | ···················· |
| W | 764 | hCoV-19/New Zealand/20CV0627/2020\|EPI_ISL_707794\|2 | ···················· | ···················· | ···················· | ···················· | ···················· | ···················· |
| W | 765 | hCoV-19/New Zealand/20CV0628/2020\|EPI_ISL_707795\|2 | ···················· | ·A·················· | ···················· | ···················· | ···················· | ··A················· |
| W | 766 | hCoV-19/New Zealand/20CV0630/2020\|EPI_ISL_707797\|2 | ···················· | ···················· | ···················· | ···················· | ···················· | ···················· |
| W | 767 | hCoV-19/New Zealand/20CV0631/2020\|EPI_ISL_707798\|2 | ···················· | ···················· | ···················· | ···················· | ···················· | ···················· |
| W | 768 | hCoV-19/New Zealand/20CV0635/2020\|EPI_ISL_707799\|2 | ···················· | ···················· | ···················· | ···················· | ···················· | ···················· |
| W | 769 | hCoV-19/USA/AZ-TG668334/2020\|EPI_ISL_696260\|2020-0 | ···················· | ···················· | ···················· | ···················· | ···················· | ···················· |
| W | 770 | hCoV-19/USA/AZ-TG668336/2020\|EPI_ISL_696261\|2020-0 | ···················· | ···················· | ···················· | ···················· | ···················· | ···················· |
| W | 771 | hCoV-19/USA/AZ-TG668340/2020\|EPI_ISL_696265\|2020-0 | ···················· | ···················· | ···················· | ···················· | ···················· | ···················· |
| W | 772 | hCoV-19/USA/AZ-TG668345/2020\|EPI_ISL_696268\|2020-0 | ···················· | ·············A······ | ···················· | ···················· | ···················· | ···················· |
| W | 773 | hCoV-19/USA/AZ-TG668350/2020\|EPI_ISL_696272\|2020-0 | ···················· | ···················· | ···················· | ···················· | ···················· | ···················· |
| W | 774 | hCoV-19/USA/AZ-TG668356/2020\|EPI_ISL_696277\|2020-0 | ···················· | ···················· | ···················· | ···················· | ···················· | ···················· |
| W | 775 | hCoV-19/USA/AZ-TG668358/2020\|EPI_ISL_696279\|2020-0 | ···················· | ···················· | ···················· | ···················· | ···················· | ···················· |
| W | 776 | hCoV-19/USA/AZ-TG668363/2020\|EPI_ISL_696283\|2020-0 | ···················· | ···················· | ···················· | ···················· | ···················· | ···················· |
| W | 777 | hCoV-19/USA/AZ-TG668367/2020\|EPI_ISL_696285\|2020-0 | ···················· | ···················· | ···················· | ···················· | ···················· | ···················· |
| W | 778 | hCoV-19/USA/AZ-TG668371/2020\|EPI_ISL_696287\|2020-0 | ···················· | ···················· | ···················· | ···················· | ···················· | ···················· |
| W | 779 | hCoV-19/USA/AZ-TG668390/2020\|EPI_ISL_696296\|2020-0 | ···················· | ···················· | ···················· | ···················· | ···················· | ···················· |
| W | 780 | hCoV-19/USA/AZ-TG668392/2020\|EPI_ISL_696298\|2020-0 | ···················· | ···················· | ···················· | ···················· | ···················· | ···················· |
| W | 781 | hCoV-19/United Arab Emirates/0679/2020\|EPI_ISL_698 | ···················· | ···················· | ···················· | ···················· | ···················· | ···················· |
| W | 782 | hCoV-19/United Arab Emirates/0680/2020\|EPI_ISL_698 | ···················· | ···················· | ···················· | ···················· | ···················· | ···················· |
| W | 783 | hCoV-19/United Arab Emirates/0681/2020\|EPI_ISL_698 | ···················· | ···················· | ···················· | ···················· | ···················· | ···················· |
| W | 784 | hCoV-19/United Arab Emirates/0682/2020\|EPI_ISL_698 | ···················· | ···················· | ···················· | ···················· | ···················· | ···················· |
| W | 785 | hCoV-19/United Arab Emirates/0685/2020\|EPI_ISL_698 | ···················· | ···················· | ···················· | ···················· | ···················· | ···················· |
| W | 786 | hCoV-19/United Arab Emirates/0686/2020\|EPI_ISL_698 | ···················· | ···················· | ···················· | ···················· | ···················· | ···················· |
| W | 787 | hCoV-19/United Arab Emirates/0687/2020\|EPI_ISL_698 | ···················· | ···················· | ···················· | ···················· | ···················· | ···················· |
| W | 788 | hCoV-19/United Arab Emirates/0688/2020\|EPI_ISL_698 | ···················· | ···················· | ···················· | ···················· | ···················· | ···················· |
| W | 789 | hCoV-19/United Arab Emirates/0689/2020\|EPI_ISL_698 | ···················· | ···················· | ···················· | ···················· | ···················· | ···················· |
| W | 790 | hCoV-19/United Arab Emirates/0690/2020\|EPI_ISL_698 | ···················· | ···················· | ···················· | ···················· | ···················· | ···················· |
| W | 791 | hCoV-19/United Arab Emirates/0691/2020\|EPI_ISL_698 | ···················· | ···················· | ···················· | ···················· | ···················· | ···················· |
| W | 792 | hCoV-19/United Arab Emirates/0696/2020\|EPI_ISL_698 | ···················· | ···················· | ···················· | ···················· | ···················· | ···················· |
| W | 793 | hCoV-19/United Arab Emirates/0697/2020\|EPI_ISL_698 | ···················· | ···················· | ···················· | ···················· | ···················· | ···················· |
| W | 794 | hCoV-19/United Arab Emirates/0803/2020\|EPI_ISL_698 | ···················· | ···················· | ···················· | ···················· | ···················· | ···················· |
| W | 795 | hCoV-19/United Arab Emirates/0804/2020\|EPI_ISL_698 | ···················· | ···················· | ···················· | ···················· | ···················· | ···················· |
| W | 796 | hCoV-19/United Arab Emirates/0805/2020\|EPI_ISL_698 | ···················· | ···················· | ···················· | ···················· | ···················· | ···················· |
| W | 797 | hCoV-19/United Arab Emirates/0806/2020\|EPI_ISL_698 | ···················· | ···················· | ···················· | ···················· | ···················· | ···················· |
| W | 798 | hCoV-19/United Arab Emirates/0807/2020\|EPI_ISL_698 | ···················· | ···················· | ···················· | ···················· | ···················· | ···················· |
| W | 799 | hCoV-19/United Arab Emirates/0808/2020\|EPI_ISL_698 | ···················· | ···················· | ···················· | ···················· | ···················· | ···················· |
| W | 800 | hCoV-19/United Arab Emirates/0809/2020\|EPI_ISL_698 | ···················· | ···················· | ···················· | ···················· | ···················· | ···················· |
| W | 801 | hCoV-19/United Arab Emirates/0810/2020\|EPI_ISL_698 | ···················· | ···················· | ···················· | ···················· | ···················· | ···················· |
| W | 802 | hCoV-19/United Arab Emirates/0811/2020\|EPI_ISL_698 | ···················· | ···················· | ···················· | ···················· | ···················· | ···················· |
| W | 803 | hCoV-19/United Arab Emirates/0812/2020\|EPI_ISL_698 | ···················· | ···················· | ···················· | ···················· | ···················· | ···················· |
| W | 804 | hCoV-19/United Arab Emirates/0813/2020\|EPI_ISL_698 | ···················· | ···················· | ···················· | ···················· | ···················· | ···················· |
| W | 805 | hCoV-19/India/UP-ACTREC-681/2020\|EPI_ISL_700306\|20 | ···················· | ·A·················· | ···················· | ···················· | ···················· | ··A················· |
| W | 806 | hCoV-19/India/MH-ACTREC-682/2020\|EPI_ISL_700307\|20 | ···················· | ···················· | ···················· | ···················· | ···················· | ···················· |
| W | 807 | hCoV-19/India/MH-ACTREC-683/2020\|EPI_ISL_700308\|20 | ···················· | ···················· | ···················· | ···················· | ···················· | ···················· |
| W | 808 | hCoV-19/India/MH-ACTREC-684/2020\|EPI_ISL_700309\|20 | ···················· | ···················· | ···················· | ···················· | ···················· | ···················· |
| W | 809 | hCoV-19/India/MH-ACTREC-685/2020\|EPI_ISL_700310\|20 | ···················· | ···················· | ···················· | ···················· | ···················· | ···················· |
| W | 810 | hCoV-19/India/WB-ACTREC-686/2020\|EPI_ISL_700311\|20 | ···················· | ···················· | ···················· | ···················· | ···················· | ···················· |
| W | 811 | hCoV-19/India/MH-ACTREC-687/2020\|EPI_ISL_700312\|20 | ···················· | ···················· | ···················· | ···················· | ···················· | ···················· |
| W | 812 | hCoV-19/India/OR-ACTREC-688/2020\|EPI_ISL_700313\|20 | ···················· | ···················· | ···················· | ···················· | ···················· | ···················· |
| W | 813 | hCoV-19/India/MH-ACTREC-689/2020\|EPI_ISL_700314\|20 | ···················· | ···················· | ···················· | ···················· | ···················· | ···················· |
| W | 814 | hCoV-19/India/MH-ACTREC-690/2020\|EPI_ISL_700315\|20 | ···················· | ···················· | ···················· | ···················· | ···················· | ···················· |
| W | 815 | hCoV-19/India/MH-ACTREC-691/2020\|EPI_ISL_700316\|20 | ···················· | ···················· | ···················· | ···················· | ···················· | ···················· |
| W | 817 | hCoV-19/India/MH-ACTREC-694/2020\|EPI_ISL_700319\|20 | ···················· | ···················· | ···················· | ···················· | ···················· | ···················· |
| W | 818 | hCoV-19/India/MH-ACTREC-695/2020\|EPI_ISL_700320\|20 | ···················· | ···················· | ···················· | ···················· | ···················· | ···················· |
| W | 819 | hCoV-19/India/MH-ACTREC-696/2020\|EPI_ISL_700321\|20 | ···················· | ·A·················· | ···················· | ···················· | ···················· | ···················· |
| W | 820 | hCoV-19/India/MH-ACTREC-697/2020\|EPI_ISL_700322\|20 | ···················· | ·A·················· | ···················· | ···················· | ···················· | ··A················· |
| W | 821 | hCoV-19/India/MH-ACTREC-698/2020\|EPI_ISL_700323\|20 | ···················· | ···················· | ···················· | ···················· | ···················· | ···················· |
| W | 823 | hCoV-19/India/MH-ACTREC-700/2020\|EPI_ISL_700325\|20 | ···················· | ···················· | ···················· | ···················· | ···················· | ···················· |
| W | 824 | hCoV-19/France/ARA-68001/2020\|EPI_ISL_700327\|2020- | ···················· | ···················· | ···················· | ···················· | ···················· | ···················· |
| W | 825 | hCoV-19/Bangladesh/CHRF-0087/2020\|EPI_ISL_700336\|2 | ···················· | ···················· | ···················· | ···················· | ···················· | ···················· |
| W | 829 | hCoV-19/Bangladesh/CHRF-0079/2020\|EPI_ISL_700342\|2 | ···················· | ···················· | ···················· | ···················· | ···················· | ···················· |
| W | 833 | hCoV-19/Bangladesh/CHRF-0086/2020\|EPI_ISL_700346\|2 | ···················· | ···················· | ···················· | ···················· | ···················· | ···················· |
| W | 835 | hCoV-19/France/ARA-SC148/2020\|EPI_ISL_700348\|2020- | ···················· | ···················· | ···················· | ···················· | ···················· | ···················· |
| W | 836 | hCoV-19/France/ARA-SC149/2020\|EPI_ISL_700349\|2020- | ···················· | ···················· | ···················· | ···················· | ···················· | ···················· |
| W | 837 | hCoV-19/France/ARA-SC155/2020\|EPI_ISL_700350\|2020- | ···················· | ···················· | ···················· | ···················· | ···················· | ···················· |
| W | 838 | hCoV-19/France/ARA-SC157/2020\|EPI_ISL_700351\|2020- | ···················· | ···················· | ···················· | ···················· | ···················· | ···················· |
| W | 839 | hCoV-19/France/ARA-SC158/2020\|EPI_ISL_700352\|2020- | ···················· | ···················· | ···················· | ···················· | ···················· | ···················· |

| | | | | | | | | |
|---|---|---|---|---|---|---|---|---|
| W | 840 | hCoV-19/France/ARA-SC164/2020\|EPI_ISL_700353\|2020- | ···················· | ···················· | ····················· | ···················· | ···················· | ···················· |
| W | 841 | hCoV-19/France/ARA-SC166/2020\|EPI_ISL_700354\|2020- | ···················· | ···················· | ····················· | ···················· | ···················· | ···················· |
| W | 842 | hCoV-19/France/ARA-SC169/2020\|EPI_ISL_700355\|2020- | ···················· | ·A·················· | ····················· | ···················· | ···················· | ···················· |
| W | 843 | hCoV-19/France/ARA-SC171/2020\|EPI_ISL_700356\|2020- | ···················· | ···················· | ····················· | ···················· | ···················· | ···················· |
| W | 844 | hCoV-19/France/ARA-SC172/2020\|EPI_ISL_700357\|2020- | ···················· | ···················· | ····················· | ···················· | ···················· | ···················· |
| W | 845 | hCoV-19/France/ARA-SC174/2020\|EPI_ISL_700358\|2020- | ···················· | ···················· | ····················· | ···················· | ···················· | ···················· |
| W | 846 | hCoV-19/France/ARA-SC181/2020\|EPI_ISL_700359\|2020- | ···················· | ···················· | ····················· | ···················· | ···················· | ···················· |
| W | 847 | hCoV-19/France/ARA-SC184/2020\|EPI_ISL_700360\|2020- | ···················· | ···················· | ····················· | ···················· | ···················· | ···················· |
| W | 848 | hCoV-19/France/ARA-SC185/2020\|EPI_ISL_700361\|2020- | ···················· | ···················· | ····················· | ···················· | ···················· | ···················· |
| W | 849 | hCoV-19/France/ARA-SC187/2020\|EPI_ISL_700362\|2020- | ···················· | ···················· | ····················· | ···················· | ···················· | ···················· |
| W | 850 | hCoV-19/France/ARA-SC188/2020\|EPI_ISL_700363\|2020- | ···················· | ···················· | ····················· | ···················· | ···················· | ···················· |
| W | 851 | hCoV-19/France/ARA-SC189/2020\|EPI_ISL_700364\|2020- | ···················· | ···················· | ····················· | ···················· | ···················· | ···················· |
| W | 852 | hCoV-19/France/ARA-SC190/2020\|EPI_ISL_700365\|2020- | ···················· | ···················· | ····················· | ···················· | ···················· | ···················· |
| W | 853 | hCoV-19/France/ARA-SC191/2020\|EPI_ISL_700366\|2020- | ···················· | ···················· | ····················· | ···················· | ···················· | ···················· |
| W | 854 | hCoV-19/France/ARA-SC193/2020\|EPI_ISL_700367\|2020- | ···················· | ···················· | ····················· | ···················· | ···················· | ···················· |
| W | 855 | hCoV-19/France/ARA-SC194/2020\|EPI_ISL_700368\|2020- | ···················· | ···················· | ····················· | ···················· | ···················· | ···················· |
| W | 856 | hCoV-19/France/ARA-SC196/2020\|EPI_ISL_700369\|2020- | ···················· | ···················· | ····················· | ···················· | ···················· | ···················· |
| W | 857 | hCoV-19/France/ARA-SC197/2020\|EPI_ISL_700370\|2020- | ···················· | ···················· | ····················· | ···················· | ···················· | ···················· |
| W | 858 | hCoV-19/France/PDL-N62441/2020\|EPI_ISL_700371\|2020 | ···················· | ···················· | ····················· | ···················· | ···················· | ···················· |
| W | 859 | hCoV-19/France/ARA-SC108/2020\|EPI_ISL_700372\|2020- | ···················· | ···················· | ····················· | ···················· | ···················· | ···················· |
| W | 860 | hCoV-19/France/ARA-SC109/2020\|EPI_ISL_700373\|2020- | ···················· | ···················· | ····················· | ···················· | ···················· | ···················· |
| W | 861 | hCoV-19/France/ARA-SC110/2020\|EPI_ISL_700374\|2020- | ···················· | ···················· | ····················· | ···················· | ···················· | ···················· |
| W | 862 | hCoV-19/France/ARA-SC111/2020\|EPI_ISL_700375\|2020- | ···················· | ·A·················· | ····················· | ···················· | ···················· | ···················· |
| W | 863 | hCoV-19/France/ARA-SC112/2020\|EPI_ISL_700376\|2020- | ···················· | ···················· | ····················· | ···················· | ···················· | ···················· |
| W | 864 | hCoV-19/France/ARA-SC113/2020\|EPI_ISL_700377\|2020- | ···················· | ···················· | ····················· | ···················· | ···················· | ···················· |
| W | 865 | hCoV-19/France/ARA-SC114/2020\|EPI_ISL_700378\|2020- | ···················· | ·A·················· | ····················· | ···················· | ···················· | ···················· |
| W | 866 | hCoV-19/France/ARA-SC115/2020\|EPI_ISL_700379\|2020- | ···················· | ···················· | ····················· | ···················· | ···················· | ···················· |
| W | 867 | hCoV-19/France/ARA-SC116/2020\|EPI_ISL_700380\|2020- | ···················· | ···················· | ····················· | ···················· | ···················· | ···················· |
| W | 868 | hCoV-19/France/ARA-SC117/2020\|EPI_ISL_700381\|2020- | ···················· | ···················· | ····················· | ···················· | ···················· | ···················· |
| W | 869 | hCoV-19/France/ARA-SC118/2020\|EPI_ISL_700382\|2020- | ···················· | ···················· | ····················· | ···················· | ···················· | ···················· |
| W | 870 | hCoV-19/France/ARA-SC119/2020\|EPI_ISL_700383\|2020- | ···················· | ···················· | ····················· | ···················· | ···················· | ···················· |
| W | 871 | hCoV-19/France/ARA-SC120/2020\|EPI_ISL_700384\|2020- | ···················· | ···················· | ····················· | ···················· | ···················· | ···················· |
| W | 872 | hCoV-19/France/ARA-SC121/2020\|EPI_ISL_700385\|2020- | ···················· | ·A·················· | ····················· | ···················· | ···················· | ···················· |
| W | 873 | hCoV-19/France/ARA-SC122/2020\|EPI_ISL_700386\|2020- | ···················· | ···················· | ····················· | ···················· | ···················· | ···················· |
| W | 874 | hCoV-19/France/ARA-SC123/2020\|EPI_ISL_700387\|2020- | ···················· | ···················· | ····················· | ···················· | ···················· | ···················· |
| W | 875 | hCoV-19/France/ARA-SC124/2020\|EPI_ISL_700388\|2020- | ···················· | ···················· | ····················· | ···················· | ···················· | ···················· |
| W | 876 | hCoV-19/France/ARA-SC125/2020\|EPI_ISL_700389\|2020- | ···················· | ···················· | ····················· | ···················· | ···················· | ···················· |
| W | 877 | hCoV-19/France/ARA-SC126/2020\|EPI_ISL_700390\|2020- | ···················· | ·A·················· | ····················· | ···················· | ···················· | ···················· |
| W | 878 | hCoV-19/France/ARA-SC130/2020\|EPI_ISL_700394\|2020- | ···················· | ···················· | ····················· | ···················· | ···················· | ···················· |
| W | 879 | hCoV-19/France/ARA-SC131/2020\|EPI_ISL_700395\|2020- | ···················· | ···················· | ····················· | ···················· | ···················· | ···················· |
| W | 880 | hCoV-19/France/ARA-SC132/2020\|EPI_ISL_700396\|2020- | ···················· | ···················· | ····················· | ···················· | ···················· | ···················· |
| W | 881 | hCoV-19/France/ARA-SC133/2020\|EPI_ISL_700397\|2020- | ···················· | ·A·················· | ····················· | ···················· | ···················· | ···················· |
| W | 882 | hCoV-19/France/ARA-SC134/2020\|EPI_ISL_700398\|2020- | ···················· | ···················· | ····················· | ···················· | ···················· | ···················· |
| W | 883 | hCoV-19/France/ARA-SC135/2020\|EPI_ISL_700399\|2020- | ···················· | ···················· | ····················· | ···················· | ···················· | ···················· |
| W | 884 | hCoV-19/Wales/ALDP-BBA082/2020\|EPI_ISL_701674\|2020 | ···················· | ···················· | ····················· | ···················· | ···················· | ···················· |
| W | 885 | hCoV-19/England/ALDP-BBB379/2020\|EPI_ISL_701676\|20 | ···················· | ···················· | ····················· | ···················· | ···················· | ···················· |
| W | 886 | hCoV-19/England/ALDP-BB54B8/2020\|EPI_ISL_701677\|20 | ···················· | ········A··········· | ····················· | ···················· | ···················· | ···················· |
| W | 887 | hCoV-19/England/ALDP-BBAFA3/2020\|EPI_ISL_701678\|20 | ···················· | ···················· | ····················· | ···················· | ···················· | ···················· |
| W | 888 | hCoV-19/England/ALDP-BBB1BB/2020\|EPI_ISL_701679\|20 | ···················· | ···················· | ····················· | ···················· | ···················· | ···················· |
| W | 889 | hCoV-19/England/ALDP-BBB036/2020\|EPI_ISL_701680\|20 | ···················· | ···················· | ····················· | ···················· | ···················· | ···················· |
| W | 890 | hCoV-19/England/ALDP-BB539D/2020\|EPI_ISL_701681\|20 | ···················· | ···················· | ····················· | ···················· | ···················· | ···················· |
| W | 891 | hCoV-19/England/ALDP-BB4FC7/2020\|EPI_ISL_701682\|20 | ···················· | ···················· | ····················· | ···················· | ···················· | ···················· |
| W | 892 | hCoV-19/England/ALDP-BB3924/2020\|EPI_ISL_701684\|20 | ···················· | ···················· | ····················· | ···················· | ···················· | ···················· |
| W | 893 | hCoV-19/England/ALDP-BBB2C7/2020\|EPI_ISL_701685\|20 | ···················· | ···················· | ····················· | ···················· | ···················· | ···················· |
| W | 894 | hCoV-19/England/ALDP-BB9F95/2020\|EPI_ISL_701686\|20 | ···················· | ···················· | ····················· | ···················· | ···················· | ···················· |
| W | 895 | hCoV-19/England/ALDP-BB51FD/2020\|EPI_ISL_701687\|20 | ···················· | ···················· | ····················· | ···················· | ···················· | ···················· |
| W | 896 | hCoV-19/England/ALDP-BB9DD7/2020\|EPI_ISL_701689\|20 | ···················· | ···················· | ····················· | ···················· | ···················· | ···················· |
| W | 897 | hCoV-19/England/ALDP-BBB081/2020\|EPI_ISL_701690\|20 | ···················· | ···················· | ····················· | ···················· | ···················· | ···················· |
| W | 898 | hCoV-19/England/ALDP-BBB151/2020\|EPI_ISL_701691\|20 | ···················· | ···················· | ····················· | ···················· | ···················· | ···················· |
| W | 899 | hCoV-19/England/ALDP-BBB0BE/2020\|EPI_ISL_701692\|20 | ···················· | ···················· | ····················· | ···················· | ···················· | ···················· |
| W | 900 | hCoV-19/England/ALDP-BB5139/2020\|EPI_ISL_701693\|20 | ···················· | ···················· | ····················· | ···················· | ···················· | ···················· |
| W | 901 | hCoV-19/England/ALDP-BB3DEC/2020\|EPI_ISL_701695\|20 | ···················· | ···················· | ····················· | ···················· | ···················· | ···················· |
| W | 902 | hCoV-19/England/ALDP-BB546D/2020\|EPI_ISL_701696\|20 | ···················· | ···················· | ····················· | ···················· | ···················· | ···················· |
| W | 903 | hCoV-19/England/ALDP-BB544F/2020\|EPI_ISL_701699\|20 | ···················· | ···················· | ····················· | ···················· | ···················· | ···················· |
| W | 904 | hCoV-19/England/ALDP-BB52AF/2020\|EPI_ISL_702940\|20 | ···················· | ···················· | ····················· | ···················· | ···················· | ···················· |
| W | 905 | hCoV-19/England/ALDP-BB5D53/2020\|EPI_ISL_702942\|20 | ···················· | ···················· | ····················· | ···················· | ···················· | ···················· |
| W | 906 | hCoV-19/England/ALDP-BB8E20/2020\|EPI_ISL_702943\|20 | ···················· | ···················· | ····················· | ···················· | ···················· | ···················· |
| W | 907 | hCoV-19/England/ALDP-BB55C4/2020\|EPI_ISL_702945\|20 | ···················· | ···················· | ····················· | ···················· | ···················· | ···················· |
| W | 908 | hCoV-19/England/ALDP-BB2C2C/2020\|EPI_ISL_702968\|20 | ···················· | ···················· | ····················· | ···················· | ···················· | ···················· |
| W | 909 | hCoV-19/England/ALDP-BB2C1D/2020\|EPI_ISL_702970\|20 | ···················· | ···················· | ····················· | ···················· | ···················· | ···················· |
| W | 910 | hCoV-19/England/ALDP-BB2B6B/2020\|EPI_ISL_702973\|20 | ···················· | ···················· | ····················· | ···················· | ···················· | ···················· |
| W | 911 | hCoV-19/England/ALDP-BB9C61/2020\|EPI_ISL_702975\|20 | ···················· | ···················· | ····················· | ···················· | ···················· | ···················· |
| W | 912 | hCoV-19/England/ALDP-BB39D9/2020\|EPI_ISL_702976\|20 | ···················· | ···················· | ····················· | ···················· | ···················· | ···················· |
| W | 913 | hCoV-19/England/ALDP-BB8EB7/2020\|EPI_ISL_702978\|20 | ···················· | ···················· | ····················· | ···················· | ···················· | ···················· |
| W | 914 | hCoV-19/England/ALDP-BB28EC/2020\|EPI_ISL_702981\|20 | ···················· | ···················· | ····················· | ···················· | ···················· | ···················· |
| W | 915 | hCoV-19/England/ALDP-BB2BD4/2020\|EPI_ISL_702983\|20 | ···················· | ···················· | ····················· | ···················· | ···················· | ···················· |
| W | 916 | hCoV-19/England/ALDP-BB9D13/2020\|EPI_ISL_702984\|20 | ···················· | ···················· | ····················· | ···················· | ···················· | ···················· |
| W | 917 | hCoV-19/England/ALDP-BB3177/2020\|EPI_ISL_702986\|20 | ···················· | ···················· | ····················· | ···················· | ···················· | ···················· |
| W | 918 | hCoV-19/England/ALDP-BB314A/2020\|EPI_ISL_702988\|20 | ···················· | ···················· | ····················· | ···················· | ···················· | ···················· |
| W | 919 | hCoV-19/England/ALDP-BB9AA3/2020\|EPI_ISL_702989\|20 | ···················· | ···················· | ····················· | ···················· | ···················· | ···················· |

| | | | | | | | | |
|---|---|---|---|---|---|---|---|---|
| W | 920 | hCoV-19/England/ALDP-BB2BB6/2020\|EPI_ISL_702991\|20 | ···················· | ···················· | ····················· | ···················· | ···················· | ···················· |
| W | 921 | hCoV-19/England/ALDP-BB3678/2020\|EPI_ISL_702993\|20 | ···················· | ···················· | ····················· | ···················· | ···················· | ···················· |
| W | 922 | hCoV-19/England/ALDP-BB3256/2020\|EPI_ISL_702995\|20 | ···················· | ···················· | ····················· | ···················· | ···················· | ············G······· |
| W | 923 | hCoV-19/England/ALDP-BB3A21/2020\|EPI_ISL_702997\|20 | ···················· | ···················· | ····················· | ···················· | ······T············· | ···················· |
| W | 924 | hCoV-19/England/ALDP-BB5D35/2020\|EPI_ISL_702998\|20 | ···················· | ···················· | ····················· | ···················· | ···················· | ···················· |
| W | 926 | hCoV-19/Wales/PHWC-4906D2/2020\|EPI_ISL_706501\|2020 | ···················· | ···················· | ····················· | ···················· | ···················· | ···················· |
| W | 931 | hCoV-19/Wales/PHWC-490748/2020\|EPI_ISL_706506\|2020 | ···················· | ···················· | ····················· | ···················· | ···················· | ···················· |
| W | 938 | hCoV-19/Wales/PHWC-4907EE/2020\|EPI_ISL_706513\|2020 | ···················· | ···················· | ····················· | ···················· | ···················· | ···················· |
| W | 939 | hCoV-19/Wales/PHWC-490818/2020\|EPI_ISL_706514\|2020 | ···················· | ···················· | ····················· | ···················· | ···················· | ···················· |
| W | 942 | hCoV-19/Wales/PHWC-490845/2020\|EPI_ISL_706517\|2020 | ···················· | ···················· | ····················· | ···················· | ···················· | ···················· |
| W | 947 | hCoV-19/Wales/PHWC-4908CD/2020\|EPI_ISL_706522\|2020 | ···················· | ···················· | ····················· | ···················· | ···················· | ···················· |
| W | 950 | hCoV-19/Wales/PHWC-4908FA/2020\|EPI_ISL_706525\|2020 | ···················· | ···················· | ····················· | ···················· | ···················· | ···················· |
| W | 956 | hCoV-19/Wales/PHWC-490C3A/2020\|EPI_ISL_706551\|2020 | ···················· | ···················· | ····················· | ···················· | ···················· | ···················· |
| W | 958 | hCoV-19/Wales/PHWC-490C58/2020\|EPI_ISL_706553\|2020 | ···················· | ···················· | ····················· | ···················· | ···················· | ···················· |
| W | 960 | hCoV-19/Wales/PHWC-490C85/2020\|EPI_ISL_706555\|2020 | ···················· | ···················· | ····················· | ···················· | ···················· | ···················· |
| W | 965 | hCoV-19/New Zealand/20CV0636/2020\|EPI_ISL_707800\|2 | ···················· | ···················· | ····················· | ···················· | ···················· | ···················· |
| W | 966 | hCoV-19/New Zealand/20CV0638/2020\|EPI_ISL_707802\|2 | ···················· | ···················· | ····················· | ···················· | ···················· | ···················· |
| W | 968 | hCoV-19/New Zealand/20CV0640/2020\|EPI_ISL_707804\|2 | ···················· | ···················· | ····················· | ···················· | ···················· | ···················· |
| W | 970 | hCoV-19/New Zealand/20CV0652/2020\|EPI_ISL_707806\|2 | ···················· | ···················· | ····················· | ···················· | ···················· | ···················· |
| W | 971 | hCoV-19/New Zealand/20CV0653/2020\|EPI_ISL_707807\|2 | ···················· | ···················· | ····················· | ···················· | ···················· | ···················· |
| W | 972 | hCoV-19/USA/MI-UM-10036766907/2020\|EPI_ISL_707813\| | ···················· | ···················· | ····················· | ···················· | ···················· | ···················· |
| W | 973 | hCoV-19/USA/MI-UM-10036794181/2020\|EPI_ISL_707814\| | ···················· | ······A······A······ | ····················· | ···················· | ···················· | ···················· |
| W | 974 | hCoV-19/USA/MI-UM-10036781165/2020\|EPI_ISL_707815\| | ···················· | ···················· | ····················· | ···················· | ···················· | ···················· |
| W | 975 | hCoV-19/USA/MI-UM-10036776666/2020\|EPI_ISL_707816\| | ···················· | ···················· | ····················· | ···················· | ···················· | ···················· |
| W | 976 | hCoV-19/USA/MI-UM-10036803260/2020\|EPI_ISL_707817\| | ···················· | ···················· | ····················· | ···················· | ···················· | ···················· |
| W | 977 | hCoV-19/USA/MI-UM-10036780471/2020\|EPI_ISL_707818\| | ···················· | ·A·················· | ····················· | ···················· | ···················· | ···················· |
| W | 979 | hCoV-19/USA/MI-UM-10036803041/2020\|EPI_ISL_707820\| | ···················· | ···················· | ····················· | ···················· | ···················· | ···················· |
| W | 981 | hCoV-19/USA/MI-UM-10036784702/2020\|EPI_ISL_707822\| | ···················· | ···················· | ····················· | ···················· | ···················· | ···················· |
| W | 982 | hCoV-19/USA/MI-UM-10036784530/2020\|EPI_ISL_707823\| | ···················· | ···················· | ····················· | ···················· | ···················· | ···················· |
| W | 983 | hCoV-19/USA/MI-UM-10036759411/2020\|EPI_ISL_707824\| | ···················· | ···················· | ····················· | ···················· | ···················· | ···················· |
| W | 984 | hCoV-19/USA/MI-UM-10036767160/2020\|EPI_ISL_707825\| | ···················· | ···················· | ····················· | ···················· | ···················· | ···················· |
| W | 985 | hCoV-19/USA/MI-UM-10036766713/2020\|EPI_ISL_707826\| | ···················· | ···················· | ····················· | ···················· | ···················· | ···················· |
| W | 986 | hCoV-19/USA/MI-UM-10036802414/2020\|EPI_ISL_707827\| | ···················· | ···················· | ····················· | ···················· | ···················· | ···················· |
| W | 987 | hCoV-19/USA/MI-UM-10036801427/2020\|EPI_ISL_707828\| | ···················· | ···················· | ····················· | ···················· | ···················· | ···················· |
| W | 988 | hCoV-19/USA/MI-UM-10036778327/2020\|EPI_ISL_707829\| | ···················· | ·············A······ | ····················· | ···················· | ···················· | ···················· |
| W | 991 | hCoV-19/USA/MI-UM-10036801411/2020\|EPI_ISL_707832\| | ···················· | ···················· | ····················· | ···················· | ···················· | ···················· |
| W | 992 | hCoV-19/USA/MI-UM-10036799861/2020\|EPI_ISL_707833\| | ···················· | ···················· | ····················· | ···················· | ···················· | ···················· |
| W | 993 | hCoV-19/USA/MI-UM-10036799095/2020\|EPI_ISL_707834\| | ···················· | ···················· | ····················· | ···················· | ···················· | ···················· |
| W | 994 | hCoV-19/USA/MI-UM-10036799267/2020\|EPI_ISL_707835\| | ···················· | ······A············· | ····················· | ···················· | ···················· | ···················· |
| W | 995 | hCoV-19/USA/MI-UM-10036801174/2020\|EPI_ISL_707836\| | ···················· | ···················· | ····················· | ···················· | ···················· | ···················· |
| W | 996 | hCoV-19/USA/MI-UM-10036795004/2020\|EPI_ISL_707837\| | ···················· | ···················· | ····················· | ···················· | ···················· | ···················· |
| W | 997 | hCoV-19/USA/MI-UM-10036797904/2020\|EPI_ISL_707838\| | ···················· | ······A············· | ····················· | ···················· | ···················· | ···················· |
| W | 998 | hCoV-19/USA/MI-UM-10036782613/2020\|EPI_ISL_707839\| | ···················· | ·A·················· | ····················· | ···················· | ···················· | ···················· |
| W | 999 | hCoV-19/USA/MI-UM-10036778374/2020\|EPI_ISL_707840\| | ···················· | ···················· | ····················· | ···················· | ···················· | ···················· |
| W | 1000 | hCoV-19/USA/MI-UM-10036797375/2020\|EPI_ISL_707841\| | ···················· | ···················· | ····················· | ···················· | ···················· | ···················· |
| W | 1001 | hCoV-19/USA/MI-UM-10036802563/2020\|EPI_ISL_707842\| | ···················· | ···················· | ····················· | ···················· | ···················· | ···················· |
| W | 1002 | hCoV-19/USA/MI-UM-10036799087/2020\|EPI_ISL_707843\| | ···················· | ···················· | ····················· | ···················· | ···················· | ···················· |
| W | 1003 | hCoV-19/USA/MI-UM-10036778541/2020\|EPI_ISL_707844\| | ···················· | ···················· | ····················· | ···················· | ···················· | ···················· |
| W | 1004 | hCoV-19/USA/MI-UM-10036782160/2020\|EPI_ISL_707845\| | ···················· | ·A·················· | ····················· | ···················· | ···················· | ···················· |
| W | 1006 | hCoV-19/USA/MI-UM-10036784170/2020\|EPI_ISL_707847\| | ···················· | ···················· | ····················· | ···················· | ···················· | ···················· |
| W | 1007 | hCoV-19/USA/MI-UM-10036798192/2020\|EPI_ISL_707848\| | ···················· | ·············A······ | ····················· | ···················· | ···················· | ···················· |
| W | 1008 | hCoV-19/USA/MI-UM-10036788119/2020\|EPI_ISL_707849\| | ···················· | ···················· | ····················· | ···················· | ···················· | ···················· |
| W | 1009 | hCoV-19/USA/MI-UM-10036782370/2020\|EPI_ISL_707850\| | ···················· | ···················· | ····················· | ···················· | ···················· | ···················· |
| W | 1010 | hCoV-19/USA/MI-UM-10036796409/2020\|EPI_ISL_707851\| | ···················· | ···················· | ····················· | ···················· | ···················· | ···················· |
| W | 1014 | hCoV-19/USA/MI-UM-10036782661/2020\|EPI_ISL_707855\| | ···················· | ···················· | ····················· | ···················· | ···················· | ···················· |
| W | 1016 | hCoV-19/USA/MI-UM-10036786992/2020\|EPI_ISL_707857\| | ···················· | ···················· | ····················· | ···················· | ···················· | ···················· |
| W | 1017 | hCoV-19/USA/MI-UM-10036789863/2020\|EPI_ISL_707858\| | ···················· | ···················· | ····················· | ···················· | ···················· | ···················· |
| W | 1018 | hCoV-19/USA/MI-UM-10036789839/2020\|EPI_ISL_707859\| | ···················· | ···················· | ····················· | ···················· | ···················· | ···················· |
| W | 1019 | hCoV-19/USA/MI-UM-10036806080/2020\|EPI_ISL_707880\| | ···················· | ···················· | ····················· | ···················· | ···················· | ···················· |
| W | 1021 | hCoV-19/USA/MI-UM-10036802137/2020\|EPI_ISL_707882\| | ···················· | ···················· | ····················· | ···················· | ···················· | ···················· |
| W | 1022 | hCoV-19/USA/CA-LACPHL-AC00004/2020\|EPI_ISL_707885\| | ···················· | ···················· | ····················· | ···················· | ···················· | ···················· |
| W | 1023 | hCoV-19/USA/CA-LACPHL-AC00005/2020\|EPI_ISL_707886\| | ···················· | ···················· | ····················· | ···················· | ···················· | ···················· |
| W | 1024 | hCoV-19/USA/CA-LACPHL-AC00007/2020\|EPI_ISL_707888\| | ···················· | ···················· | ····················· | ···················· | ···················· | ···················· |
| W | 1025 | hCoV-19/USA/AZ-TG666122/2020\|EPI_ISL_696208\|2020-0 | ···················· | ···················· | ····················· | ···················· | ···················· | ···················· |
| W | 1026 | hCoV-19/USA/AZ-TG666123/2020\|EPI_ISL_696209\|2020-0 | ···················· | ···················· | ····················· | ···················· | ···················· | ···················· |
| W | 1027 | hCoV-19/USA/AZ-TG666124/2020\|EPI_ISL_696210\|2020-0 | ···················· | ···················· | ····················· | ···················· | ···················· | ···················· |
| W | 1028 | hCoV-19/USA/AZ-TG666128/2020\|EPI_ISL_696214\|2020-0 | ···················· | ···················· | ····················· | ···················· | ···················· | ···················· |
| W | 1029 | hCoV-19/USA/AZ-TG666131/2020\|EPI_ISL_696216\|2020-0 | ···················· | ···················· | ····················· | ···················· | ···················· | ···················· |
| W | 1030 | hCoV-19/USA/AZ-TG666135/2020\|EPI_ISL_696219\|2020-0 | ···················· | ···················· | ····················· | ···················· | ···················· | ···················· |
| W | 1031 | hCoV-19/USA/AZ-TG666140/2020\|EPI_ISL_696223\|2020-0 | ···················· | ···················· | ····················· | ···················· | ···················· | ···················· |
| W | 1032 | hCoV-19/USA/AZ-TG666141/2020\|EPI_ISL_696224\|2020-0 | ···················· | ···················· | ····················· | ···················· | ···················· | ···················· |
| W | 1033 | hCoV-19/USA/AZ-TG666143/2020\|EPI_ISL_696226\|2020-0 | ···················· | ···················· | ····················· | ···················· | ···················· | ···················· |
| W | 1034 | hCoV-19/USA/AZ-TG666148/2020\|EPI_ISL_696229\|2020-0 | ···················· | ···················· | ····················· | ···················· | ···················· | ···················· |
| W | 1035 | hCoV-19/USA/AZ-TG666149/2020\|EPI_ISL_696230\|2020-0 | ···················· | ···················· | ····················· | ···················· | ···················· | ···················· |
| W | 1036 | hCoV-19/USA/AZ-TG666152/2020\|EPI_ISL_696233\|2020-0 | ···················· | ···················· | ····················· | ···················· | ···················· | ···················· |
| W | 1037 | hCoV-19/USA/AZ-TG666156/2020\|EPI_ISL_696237\|2020-0 | ···················· | ···················· | ····················· | ···················· | ···················· | ···················· |
| W | 1038 | hCoV-19/USA/AZ-TG666157/2020\|EPI_ISL_696238\|2020-0 | ···················· | ···················· | ····················· | ···················· | ···················· | ···················· |
| W | 1039 | hCoV-19/USA/AZ-TG666160/2020\|EPI_ISL_696241\|2020-0 | ···················· | ···················· | ····················· | ···················· | ···················· | ···················· |
| W | 1040 | hCoV-19/USA/AZ-TG666162/2020\|EPI_ISL_696242\|2020-0 | ···················· | ···················· | ····················· | ···················· | ···················· | ···················· |
| W | 1041 | hCoV-19/USA/AZ-TG666165/2020\|EPI_ISL_696245\|2020-0 | ···················· | ···················· | ····················· | ···················· | ···················· | ···················· |

| | | | | | | | | |
|---|---|---|---|---|---|---|---|---|
| W | 1042 | hCoV-19/USA/AZ-TG666168/2020\|EPI_ISL_696248\|2020-0 | .................... | .................... | ..................... | .................... | .................... | .................... |
| W | 1043 | hCoV-19/USA/AZ-TG666169/2020\|EPI_ISL_696249\|2020-0 | .................... | .................... | ..................... | .................... | .................... | .................... |
| W | 1044 | hCoV-19/USA/AZ-TG666172/2020\|EPI_ISL_696251\|2020-0 | .................... | .................... | ..................... | .................... | .................... | .................... |
| W | 1045 | hCoV-19/USA/AZ-TG666174/2020\|EPI_ISL_696252\|2020-0 | .................... | .................... | ..................... | .................... | .................... | .................... |
| W | 1046 | hCoV-19/USA/AZ-TG666175/2020\|EPI_ISL_696253\|2020-0 | .................... | .................... | ..................... | .................... | .................... | .................... |
| W | 1047 | hCoV-19/USA/AZ-TG666177/2020\|EPI_ISL_696254\|2020-0 | .................... | .................... | ..................... | .................... | .................... | .................... |
| W | 1048 | hCoV-19/USA/AZ-TG668327/2020\|EPI_ISL_696256\|2020-0 | .................... | .................... | ..................... | .................... | .................... | .................... |
| W | 1049 | hCoV-19/United Arab Emirates/0644/2020\|EPI_ISL_698 | .................... | .................... | ..................... | .................... | .................... | .................... |
| W | 1050 | hCoV-19/United Arab Emirates/0645/2020\|EPI_ISL_698 | .................... | .................... | ..................... | .................... | .................... | .................... |
| W | 1051 | hCoV-19/United Arab Emirates/0646/2020\|EPI_ISL_698 | .................... | .................... | ..................... | .................... | .................... | .................... |
| W | 1052 | hCoV-19/United Arab Emirates/0647/2020\|EPI_ISL_698 | .................... | .................... | ..................... | .................... | .................... | .................... |
| W | 1053 | hCoV-19/United Arab Emirates/0648/2020\|EPI_ISL_698 | .................... | .................... | ..................... | .................... | .................... | .................... |
| W | 1054 | hCoV-19/United Arab Emirates/0649/2020\|EPI_ISL_698 | .................... | .................... | ..................... | .................... | .................... | .................... |
| W | 1055 | hCoV-19/United Arab Emirates/0650/2020\|EPI_ISL_698 | .................... | .................... | ..................... | .................... | .................... | .................... |
| W | 1056 | hCoV-19/United Arab Emirates/0651/2020\|EPI_ISL_698 | .................... | .................... | ..................... | .................... | .................... | .................... |
| W | 1057 | hCoV-19/United Arab Emirates/0653/2020\|EPI_ISL_698 | .................... | .A.................. | ..................... | .................... | .................... | .................... |
| W | 1058 | hCoV-19/United Arab Emirates/0654/2020\|EPI_ISL_698 | .................... | .................... | ..................... | .................... | .................... | .................... |
| W | 1059 | hCoV-19/United Arab Emirates/0655/2020\|EPI_ISL_698 | .................... | .................... | ..................... | .................... | .................... | .................... |
| W | 1060 | hCoV-19/United Arab Emirates/0656/2020\|EPI_ISL_698 | .................... | .................... | ..................... | .................... | .................... | .................... |
| W | 1061 | hCoV-19/United Arab Emirates/0657/2020\|EPI_ISL_698 | .................... | .................... | ..................... | .................... | .................... | .................... |
| W | 1062 | hCoV-19/United Arab Emirates/0659/2020\|EPI_ISL_698 | .................... | .................... | ..................... | .................... | .................... | .................... |
| W | 1063 | hCoV-19/United Arab Emirates/0660/2020\|EPI_ISL_698 | .................... | .................... | ..................... | .................... | .................... | .................... |
| W | 1064 | hCoV-19/United Arab Emirates/0661/2020\|EPI_ISL_698 | .................... | .................... | ..................... | .................... | .................... | .................... |
| W | 1065 | hCoV-19/United Arab Emirates/0662/2020\|EPI_ISL_698 | .................... | .................... | ..................... | .................... | .................... | .................... |
| W | 1066 | hCoV-19/United Arab Emirates/0663/2020\|EPI_ISL_698 | .................... | .................... | ..................... | .................... | .................... | .................... |
| W | 1067 | hCoV-19/United Arab Emirates/0664/2020\|EPI_ISL_698 | .................... | .................... | ..................... | .................... | .................... | .................... |
| W | 1068 | hCoV-19/United Arab Emirates/0665/2020\|EPI_ISL_698 | .................... | .................... | ..................... | .................... | .................... | .................... |
| W | 1069 | hCoV-19/United Arab Emirates/0666/2020\|EPI_ISL_698 | .................... | .................... | ..................... | .................... | .................... | .................... |
| W | 1070 | hCoV-19/United Arab Emirates/0668/2020\|EPI_ISL_698 | .................... | .................... | ..................... | .................... | .................... | .................... |
| W | 1071 | hCoV-19/United Arab Emirates/0669/2020\|EPI_ISL_698 | .................... | .................... | ..................... | .................... | .................... | .................... |
| W | 1072 | hCoV-19/United Arab Emirates/0670/2020\|EPI_ISL_698 | .................... | .................... | ..................... | .................... | .................... | .................... |
| W | 1073 | hCoV-19/United Arab Emirates/0671/2020\|EPI_ISL_698 | .................... | .................... | ..................... | .................... | .................... | .................... |
| W | 1074 | hCoV-19/United Arab Emirates/0672/2020\|EPI_ISL_698 | .................... | .................... | ..................... | .................... | .................... | .................... |
| W | 1075 | hCoV-19/United Arab Emirates/0673/2020\|EPI_ISL_698 | .................... | .................... | ..................... | .................... | .................... | .................... |
| W | 1076 | hCoV-19/United Arab Emirates/0674/2020\|EPI_ISL_698 | .................... | .................... | ..................... | .................... | .................... | .................... |
| W | 1077 | hCoV-19/United Arab Emirates/0675/2020\|EPI_ISL_698 | .................... | .................... | ..................... | .................... | .................... | .................... |
| W | 1078 | hCoV-19/United Arab Emirates/0676/2020\|EPI_ISL_698 | .................... | .................... | ..................... | .................... | .................... | .................... |
| W | 1079 | hCoV-19/United Arab Emirates/0677/2020\|EPI_ISL_698 | .................... | .................... | ..................... | .................... | .................... | .................... |
| W | 1080 | hCoV-19/United Arab Emirates/0678/2020\|EPI_ISL_698 | .................... | .................... | ..................... | .................... | .................... | .................... |
| W | 1081 | hCoV-19/United Arab Emirates/0692/2020\|EPI_ISL_698 | .................... | .................... | ..................... | .................... | .................... | .................... |
| W | 1082 | hCoV-19/United Arab Emirates/0693/2020\|EPI_ISL_698 | .................... | .................... | ..................... | .................... | .................... | .................... |
| W | 1083 | hCoV-19/United Arab Emirates/0694/2020\|EPI_ISL_698 | .................... | .................... | ..................... | .................... | .................... | .................... |
| W | 1084 | hCoV-19/United Arab Emirates/0695/2020\|EPI_ISL_698 | .................... | .................... | ..................... | .................... | .................... | .................... |
| W | 1085 | hCoV-19/United Arab Emirates/0698/2020\|EPI_ISL_698 | .................... | .................... | ..................... | .................... | .................... | .................... |
| W | 1086 | hCoV-19/United Arab Emirates/0699/2020\|EPI_ISL_698 | .................... | .................... | ..................... | .................... | .................... | .................... |
| W | 1087 | hCoV-19/United Arab Emirates/0700/2020\|EPI_ISL_698 | .................... | .................... | ..................... | .................... | .................... | .................... |
| W | 1088 | hCoV-19/United Arab Emirates/0701/2020\|EPI_ISL_698 | .................... | .................... | ..................... | .................... | .................... | .................... |
| W | 1089 | hCoV-19/United Arab Emirates/0702/2020\|EPI_ISL_698 | .................... | .................... | ..................... | .................... | .................... | .................... |
| W | 1090 | hCoV-19/United Arab Emirates/0703/2020\|EPI_ISL_698 | .................... | .................... | ..................... | .................... | .................... | .................... |
| W | 1091 | hCoV-19/United Arab Emirates/0704/2020\|EPI_ISL_698 | .................... | .................... | ..................... | .................... | .................... | .................... |
| W | 1092 | hCoV-19/United Arab Emirates/0705/2020\|EPI_ISL_698 | .................... | .................... | ..................... | .................... | .................... | .................... |
| W | 1093 | hCoV-19/United Arab Emirates/0706/2020\|EPI_ISL_698 | .................... | .................... | ..................... | .................... | .................... | .................... |
| W | 1094 | hCoV-19/United Arab Emirates/0707/2020\|EPI_ISL_698 | .................... | .................... | ..................... | .................... | .................... | .................... |
| W | 1095 | hCoV-19/United Arab Emirates/0708/2020\|EPI_ISL_698 | .................... | .................... | ..................... | .................... | .................... | .................... |
| W | 1096 | hCoV-19/United Arab Emirates/0709/2020\|EPI_ISL_698 | .................... | .................... | ..................... | .................... | .................... | .................... |
| W | 1097 | hCoV-19/United Arab Emirates/0710/2020\|EPI_ISL_698 | .................... | .................... | ..................... | .................... | .................... | .................... |
| W | 1098 | hCoV-19/United Arab Emirates/0711/2020\|EPI_ISL_698 | .................... | .................... | ..................... | .................... | .................... | .................... |
| W | 1099 | hCoV-19/United Arab Emirates/0712/2020\|EPI_ISL_698 | .................... | .................... | ..................... | .................... | .................... | .................... |
| W | 1100 | hCoV-19/United Arab Emirates/0713/2020\|EPI_ISL_698 | .................... | .................... | ..................... | .................... | .................... | .................... |
| W | 1101 | hCoV-19/United Arab Emirates/0714/2020\|EPI_ISL_698 | .................... | .................... | ..................... | .................... | .................... | .................... |
| W | 1102 | hCoV-19/United Arab Emirates/0715/2020\|EPI_ISL_698 | .................... | .................... | ..................... | .................... | .................... | .................... |
| W | 1103 | hCoV-19/United Arab Emirates/0716/2020\|EPI_ISL_698 | .................... | .................... | ..................... | .................... | .................... | .................... |
| W | 1104 | hCoV-19/United Arab Emirates/0717/2020\|EPI_ISL_698 | .................... | .................... | ..................... | .................... | .................... | .................... |
| W | 1105 | hCoV-19/United Arab Emirates/0718/2020\|EPI_ISL_698 | .................... | .................... | ..................... | .................... | .................... | .................... |
| W | 1106 | hCoV-19/United Arab Emirates/0719/2020\|EPI_ISL_698 | .................... | .................... | ..................... | .................... | .................... | .................... |
| W | 1107 | hCoV-19/United Arab Emirates/0720/2020\|EPI_ISL_698 | .................... | .................... | ..................... | .................... | .................... | .................... |
| W | 1108 | hCoV-19/United Arab Emirates/0721/2020\|EPI_ISL_698 | .................... | .................... | ..................... | .................... | .................... | .................... |
| W | 1109 | hCoV-19/United Arab Emirates/0722/2020\|EPI_ISL_698 | .................... | .................... | ..................... | .................... | .................... | .................... |
| W | 1110 | hCoV-19/United Arab Emirates/0723/2020\|EPI_ISL_698 | .................... | .................... | ..................... | .................... | .................... | .................... |
| W | 1111 | hCoV-19/United Arab Emirates/0724/2020\|EPI_ISL_698 | .................... | .................... | ..................... | .................... | .................... | .................... |
| W | 1112 | hCoV-19/United Arab Emirates/0725/2020\|EPI_ISL_698 | .................... | .................... | ..................... | .................... | .................... | .................... |
| W | 1113 | hCoV-19/United Arab Emirates/0726/2020\|EPI_ISL_698 | .................... | .................... | ..................... | .................... | .................... | .................... |
| W | 1114 | hCoV-19/United Arab Emirates/0727/2020\|EPI_ISL_698 | .................... | .................... | ..................... | .................... | .................... | .................... |
| W | 1115 | hCoV-19/United Arab Emirates/0728/2020\|EPI_ISL_698 | .................... | .................... | ..................... | .................... | .................... | .................... |
| W | 1116 | hCoV-19/United Arab Emirates/0729/2020\|EPI_ISL_698 | .................... | .................... | ..................... | .................... | .................... | .................... |
| W | 1117 | hCoV-19/United Arab Emirates/0730/2020\|EPI_ISL_698 | .................... | .................... | ..................... | .................... | .................... | .................... |
| W | 1118 | hCoV-19/United Arab Emirates/0731/2020\|EPI_ISL_698 | .................... | .................... | ..................... | .................... | .................... | .................... |
| W | 1119 | hCoV-19/United Arab Emirates/0732/2020\|EPI_ISL_698 | .................... | .................... | ..................... | .................... | .................... | .................... |
| W | 1120 | hCoV-19/United Arab Emirates/0734/2020\|EPI_ISL_698 | .................... | .................... | ..................... | .................... | .................... | .................... |
| W | 1121 | hCoV-19/United Arab Emirates/0735/2020\|EPI_ISL_698 | .................... | .................... | ..................... | .................... | .................... | .................... |

| | | | | | | | | |
|---|---|---|---|---|---|---|---|---|
| W | 1122 | hCoV-19/United Arab Emirates/0737/2020\|EPI_ISL_698 | ···················· | ···················· | ···················· | ···················· | ···················· | ···················· |
| W | 1123 | hCoV-19/United Arab Emirates/0738/2020\|EPI_ISL_698 | ···················· | ···················· | ···················· | ···················· | ···················· | ···················· |
| W | 1124 | hCoV-19/United Arab Emirates/0739/2020\|EPI_ISL_698 | ···················· | ···················· | ···················· | ···················· | ···················· | ···················· |
| W | 1125 | hCoV-19/United Arab Emirates/0740/2020\|EPI_ISL_698 | ···················· | ···················· | ···················· | ···················· | ···················· | ···················· |
| W | 1126 | hCoV-19/United Arab Emirates/0741/2020\|EPI_ISL_698 | ···················· | ···················· | ···················· | ···················· | ···················· | ···················· |
| W | 1127 | hCoV-19/United Arab Emirates/0742/2020\|EPI_ISL_698 | ···················· | ···················· | ···················· | ···················· | ···················· | ···················· |
| W | 1128 | hCoV-19/United Arab Emirates/0743/2020\|EPI_ISL_698 | ···················· | ···················· | ···················· | ···················· | ···················· | ···················· |
| W | 1129 | hCoV-19/United Arab Emirates/0744/2020\|EPI_ISL_698 | ···················· | ···················· | ···················· | ···················· | ···················· | ···················· |
| W | 1130 | hCoV-19/United Arab Emirates/0745/2020\|EPI_ISL_698 | ···················· | ···················· | ···················· | ···················· | ···················· | ···················· |
| W | 1131 | hCoV-19/United Arab Emirates/0746/2020\|EPI_ISL_698 | ···················· | ···················· | ···················· | ···················· | ···················· | ···················· |
| W | 1132 | hCoV-19/United Arab Emirates/0747/2020\|EPI_ISL_698 | ···················· | ···················· | ···················· | ···················· | ···················· | ···················· |
| W | 1133 | hCoV-19/United Arab Emirates/0748/2020\|EPI_ISL_698 | ···················· | ···················· | ···················· | ···················· | ···················· | ···················· |
| W | 1134 | hCoV-19/United Arab Emirates/0749/2020\|EPI_ISL_698 | ···················· | ···················· | ···················· | ···················· | ···················· | ···················· |
| W | 1135 | hCoV-19/United Arab Emirates/0750/2020\|EPI_ISL_698 | ···················· | ···················· | ···················· | ···················· | ···················· | ···················· |
| W | 1136 | hCoV-19/France/ARA-SC137/2020\|EPI_ISL_700401\|2020- | ···················· | ···················· | ···················· | ···················· | ···················· | ···················· |
| W | 1137 | hCoV-19/France/ARA-SC138/2020\|EPI_ISL_700402\|2020- | ···················· | ···················· | ···················· | ···················· | ···················· | ···················· |
| W | 1138 | hCoV-19/France/ARA-SC139/2020\|EPI_ISL_700403\|2020- | ···················· | ·A·················· | ···················· | ···················· | ···················· | ···················· |
| W | 1139 | hCoV-19/France/ARA-SC140/2020\|EPI_ISL_700404\|2020- | ···················· | ···················· | ···················· | ···················· | ···················· | ···················· |
| W | 1140 | hCoV-19/France/ARA-SC143/2020\|EPI_ISL_700407\|2020- | ···················· | ···················· | ···················· | ···················· | ···················· | ···················· |
| W | 1142 | hCoV-19/France/ARA-SC145/2020\|EPI_ISL_700409\|2020- | ···················· | ·A·················· | ···················· | ···················· | ···················· | ···················· |
| W | 1143 | hCoV-19/France/ARA-SC146/2020\|EPI_ISL_700410\|2020- | ···················· | ·A·················· | ···················· | ···················· | ···················· | ···················· |
| W | 1149 | hCoV-19/England/ALDP-BB37EE/2020\|EPI_ISL_701701\|20 | ···················· | ···················· | ···················· | ···················· | ···················· | ···················· |
| W | 1150 | hCoV-19/England/ALDP-BBB26D/2020\|EPI_ISL_701703\|20 | ···················· | ···················· | ···················· | ···················· | ···················· | ···················· |
| W | 1151 | hCoV-19/England/ALDP-BB53E8/2020\|EPI_ISL_701704\|20 | ···················· | ···················· | ···················· | ···················· | ···················· | ···················· |
| W | 1152 | hCoV-19/England/ALDP-BB5421/2020\|EPI_ISL_701705\|20 | ···················· | ···················· | ···················· | ···················· | ···················· | ···················· |
| W | 1153 | hCoV-19/England/ALDP-BBA732/2020\|EPI_ISL_701707\|20 | ···················· | ···················· | ···················· | ···················· | ······T············· | ···················· |
| W | 1154 | hCoV-19/England/ALDP-BBA37A/2020\|EPI_ISL_701708\|20 | ···················· | ···················· | ···················· | ···················· | ···················· | ···················· |
| W | 1155 | hCoV-19/England/ALDP-BB5351/2020\|EPI_ISL_701709\|20 | ···················· | ···················· | ···················· | ···················· | ···················· | ···················· |
| W | 1156 | hCoV-19/England/ALDP-BBA18F/2020\|EPI_ISL_701710\|20 | ···················· | ····A··············· | ···················· | ···················· | ···················· | ···················· |
| W | 1157 | hCoV-19/England/ALDP-BB51C0/2020\|EPI_ISL_701711\|20 | ···················· | ···················· | ···················· | ···················· | ···················· | ···················· |
| W | 1158 | hCoV-19/England/ALDP-BBB2E5/2020\|EPI_ISL_701712\|20 | ···················· | ···················· | ···················· | ···················· | ···················· | ···················· |
| W | 1159 | hCoV-19/England/ALDP-BB3F6E/2020\|EPI_ISL_701713\|20 | ···················· | ···················· | ···················· | ···················· | ···················· | ···················· |
| W | 1160 | hCoV-19/England/ALDP-BB3793/2020\|EPI_ISL_701714\|20 | ···················· | ···················· | ···················· | ···················· | ···················· | ···················· |
| W | 1161 | hCoV-19/England/ALDP-BB3CD0/2020\|EPI_ISL_701715\|20 | ···················· | ···················· | ···················· | ···················· | ······T············· | ···················· |
| W | 1162 | hCoV-19/England/ALDP-BB5263/2020\|EPI_ISL_701716\|20 | ···················· | ···················· | ···················· | ···················· | ···················· | ···················· |
| W | 1163 | hCoV-19/England/ALDP-BBA064/2020\|EPI_ISL_701718\|20 | ···················· | ···················· | ···················· | ···················· | ···················· | ···················· |
| W | 1164 | hCoV-19/England/ALDP-BBB35B/2020\|EPI_ISL_701719\|20 | ···················· | ···················· | ···················· | ···················· | ···················· | ···················· |
| W | 1165 | hCoV-19/England/ALDP-BBB0CD/2020\|EPI_ISL_701721\|20 | ···················· | ···················· | ···················· | ···················· | ···················· | ···················· |
| W | 1166 | hCoV-19/England/ALDP-BBAC33/2020\|EPI_ISL_701723\|20 | ···················· | ···················· | ···················· | ···················· | ···················· | ···················· |
| W | 1167 | hCoV-19/England/ALDP-BB54F4/2020\|EPI_ISL_701725\|20 | ···················· | ···················· | ···················· | ···················· | ···················· | ···················· |
| W | 1168 | hCoV-19/England/ALDP-BB3F7D/2020\|EPI_ISL_701726\|20 | ···················· | ···················· | ···················· | ···················· | ···················· | ···················· |
| W | 1169 | hCoV-19/England/ALDP-BBB449/2020\|EPI_ISL_701727\|20 | ···················· | ···················· | ···················· | ···················· | ···················· | ···················· |
| W | 1170 | hCoV-19/England/ALDP-BBB4EF/2020\|EPI_ISL_701728\|20 | ···················· | ···················· | ···················· | ···················· | ···················· | ···················· |
| W | 1171 | hCoV-19/England/ALDP-BB3784/2020\|EPI_ISL_701730\|20 | ···················· | ···················· | ···················· | ···················· | ···················· | ···················· |
| W | 1172 | hCoV-19/England/ALDP-BB5193/2020\|EPI_ISL_701731\|20 | ···················· | ···················· | ···················· | ···················· | ···················· | ···················· |
| W | 1173 | hCoV-19/England/ALDP-BB3DDD/2020\|EPI_ISL_701732\|20 | ···················· | ···················· | ···················· | ···················· | ···················· | ···················· |
| W | 1174 | hCoV-19/England/ALDP-BBB124/2020\|EPI_ISL_701733\|20 | ···················· | ···················· | ···················· | ···················· | ···················· | ···················· |
| W | 1175 | hCoV-19/England/ALDP-BB510C/2020\|EPI_ISL_701734\|20 | ···················· | ···················· | ···················· | ···················· | ···················· | ···················· |
| W | 1176 | hCoV-19/England/ALDP-BB9DE6/2020\|EPI_ISL_701736\|20 | ···················· | ···················· | ···················· | ···················· | ···················· | ···················· |
| W | 1177 | hCoV-19/England/ALDP-BBB485/2020\|EPI_ISL_701738\|20 | ···················· | ···················· | ···················· | ···················· | ···················· | ···················· |
| W | 1178 | hCoV-19/England/ALDP-BB3D37/2020\|EPI_ISL_701739\|20 | ···················· | ···················· | ···················· | ···················· | ···················· | ···················· |
| W | 1179 | hCoV-19/England/ALDP-BB5069/2020\|EPI_ISL_701740\|20 | ···················· | ·A·················· | ···················· | ···················· | ···················· | ···················· |
| W | 1180 | hCoV-19/England/ALDP-BB560D/2020\|EPI_ISL_701741\|20 | ···················· | ·········T·········· | ···················· | ···················· | ···················· | ···················· |
| W | 1181 | hCoV-19/England/ALDP-BB9FC2/2020\|EPI_ISL_701742\|20 | ···················· | ···················· | ···················· | ···················· | ···················· | ···················· |
| W | 1182 | hCoV-19/England/ALDP-BB54C7/2020\|EPI_ISL_701745\|20 | ···················· | ···················· | ···················· | ···················· | ···················· | ···················· |
| W | 1183 | hCoV-19/England/ALDP-BB3CC1/2020\|EPI_ISL_701746\|20 | ···················· | ···················· | ···················· | ···················· | ···················· | ···················· |
| W | 1184 | hCoV-19/England/ALDP-BBB203/2020\|EPI_ISL_701747\|20 | ···················· | ···················· | ···················· | ···················· | ···················· | ···················· |
| W | 1185 | hCoV-19/England/ALDP-BBA88A/2020\|EPI_ISL_701748\|20 | ···················· | ···················· | ···················· | ···················· | ···················· | ···················· |
| W | 1186 | hCoV-19/England/ALDP-BB3836/2020\|EPI_ISL_701749\|20 | ···················· | ···················· | ···················· | ···················· | ···················· | ···················· |
| W | 1187 | hCoV-19/England/ALDP-BB9E3E/2020\|EPI_ISL_701750\|20 | ···················· | ···················· | ···················· | ···················· | ···················· | ···················· |
| W | 1188 | hCoV-19/England/ALDP-BBAC51/2020\|EPI_ISL_701752\|20 | ···················· | ···················· | ···················· | ···················· | ···················· | ···················· |
| W | 1189 | hCoV-19/Wales/ALDP-BB55F1/2020\|EPI_ISL_701753\|2020 | ···················· | ···················· | ···················· | ···················· | ···················· | ···················· |
| W | 1190 | hCoV-19/England/ALDP-BB3AF4/2020\|EPI_ISL_701754\|20 | ···················· | ···················· | ···················· | ···················· | ···················· | ···················· |
| W | 1191 | hCoV-19/England/ALDP-BBB1E8/2020\|EPI_ISL_701756\|20 | ···················· | ···················· | ···················· | ···················· | ···················· | ···················· |
| W | 1192 | hCoV-19/England/MILK-BAB5A3/2020\|EPI_ISL_705200\|20 | ···················· | ···················· | ···················· | ···················· | ···················· | ···················· |
| W | 1193 | hCoV-19/England/MILK-BAAA0F/2020\|EPI_ISL_705202\|20 | ···················· | ···················· | ···················· | ···················· | ···················· | ···················· |
| W | 1197 | hCoV-19/Wales/PHWC-490A8B/2020\|EPI_ISL_706533\|2020 | ···················· | ···················· | ···················· | ···················· | ···················· | ···················· |
| W | 1198 | hCoV-19/Wales/PHWC-490AA9/2020\|EPI_ISL_706534\|2020 | ···················· | ···················· | ···················· | ···················· | ···················· | ···················· |
| W | 1200 | hCoV-19/Wales/PHWC-490AD6/2020\|EPI_ISL_706536\|2020 | ···················· | ···················· | ···················· | ···················· | ···················· | ···················· |
| W | 1201 | hCoV-19/Wales/PHWC-490B00/2020\|EPI_ISL_706537\|2020 | ···················· | ···················· | ···················· | ···················· | ···A················ | ···················· |
| W | 1205 | hCoV-19/Wales/PHWC-490B5B/2020\|EPI_ISL_706541\|2020 | ···················· | ···················· | ···················· | ···················· | ···················· | ···················· |
| W | 1208 | hCoV-19/Wales/PHWC-490BC4/2020\|EPI_ISL_706544\|2020 | ···················· | ···················· | ···················· | ···················· | ···················· | ···················· |
| W | 1210 | hCoV-19/Wales/PHWC-490BE2/2020\|EPI_ISL_706546\|2020 | ···················· | ···················· | ···················· | ···················· | ···················· | ···················· |
| W | 1211 | hCoV-19/Wales/PHWC-490BF1/2020\|EPI_ISL_706547\|2020 | ···················· | ···················· | ···················· | ···················· | ···················· | ···················· |
| W | 1212 | hCoV-19/Wales/PHWC-490C0D/2020\|EPI_ISL_706548\|2020 | ···················· | ···················· | ···················· | ···················· | ···················· | ···················· |
| W | 1213 | hCoV-19/Wales/PHWC-490C1C/2020\|EPI_ISL_706549\|2020 | ···················· | ···················· | ···················· | ···················· | ···················· | ···················· |
| W | 1218 | hCoV-19/Wales/PHWC-490D28/2020\|EPI_ISL_706564\|2020 | ···················· | ···················· | ···················· | ···················· | ···················· | ···················· |
| W | 1221 | hCoV-19/Wales/PHWC-490D55/2020\|EPI_ISL_706567\|2020 | ···················· | ···················· | ···················· | ···················· | ···················· | ···················· |
| W | 1226 | hCoV-19/Wales/PHWC-490DDD/2020\|EPI_ISL_706572\|2020 | ···················· | ···················· | ···················· | ···················· | ···················· | ···················· |
| W | 1230 | hCoV-19/Wales/PHWC-490E34/2020\|EPI_ISL_706576\|2020 | ···················· | ···················· | ···················· | ···················· | ···················· | ···················· |

| | | | | | | | | |
|---|---|---|---|---|---|---|---|---|
| W | 1233 | hCoV-19/Wales/PHWC-490E61/2020\|EPI_ISL_706579\|2020 | ···················· | ···················· | ···················· | ···················· | ···················· | ···················· |
| W | 1234 | hCoV-19/Wales/PHWC-490E8F/2020\|EPI_ISL_706580\|2020 | ···················· | ···················· | ···················· | ···················· | ···················· | ···················· |
| W | 1235 | hCoV-19/Wales/PHWC-490E9E/2020\|EPI_ISL_706581\|2020 | ···················· | ···················· | ···················· | ···················· | ···················· | ···················· |
| W | 1236 | hCoV-19/Wales/PHWC-490EBC/2020\|EPI_ISL_706582\|2020 | ···················· | ···················· | ···················· | ···················· | ···················· | ···················· |
| W | 1237 | hCoV-19/Wales/PHWC-490ECB/2020\|EPI_ISL_706583\|2020 | ···················· | ···················· | ···················· | ···················· | ···················· | ···················· |
| W | 1238 | hCoV-19/Wales/PHWC-490EE9/2020\|EPI_ISL_706584\|2020 | ···················· | ···················· | ···················· | ···················· | ···················· | ···················· |
| W | 1239 | hCoV-19/Wales/PHWC-490EF8/2020\|EPI_ISL_706585\|2020 | ···················· | ···················· | ···················· | ···················· | ···················· | ···················· |
| W | 1240 | hCoV-19/Wales/PHWC-490F13/2020\|EPI_ISL_706586\|2020 | ···················· | ···················· | ···················· | ···················· | ···················· | ···················· |
| W | 1242 | hCoV-19/Wales/PHWC-490F31/2020\|EPI_ISL_706588\|2020 | ···················· | ···················· | ···················· | ···················· | ···················· | ···················· |
| W | 1243 | hCoV-19/Wales/PHWC-490F5F/2020\|EPI_ISL_706589\|2020 | ···················· | ···················· | ···················· | ···················· | ···················· | ···················· |
| W | 1244 | hCoV-19/Wales/PHWC-490F6E/2020\|EPI_ISL_706590\|2020 | ···················· | ···················· | ···················· | ···················· | ···················· | ···················· |
| W | 1245 | hCoV-19/Wales/PHWC-490F7D/2020\|EPI_ISL_706591\|2020 | ···················· | ···················· | ···················· | ···················· | ···················· | ···················· |
| W | 1246 | hCoV-19/Wales/PHWC-490F8C/2020\|EPI_ISL_706592\|2020 | ···················· | ···················· | ···················· | ···················· | ···················· | ···················· |
| W | 1247 | hCoV-19/Wales/PHWC-490F9B/2020\|EPI_ISL_706593\|2020 | ···················· | ···················· | ···················· | ···················· | ···················· | ···················· |
| W | 1248 | hCoV-19/Wales/PHWC-490FAA/2020\|EPI_ISL_706594\|2020 | ···················· | ···················· | ···················· | ···················· | ···················· | ···················· |
| W | 1249 | hCoV-19/Wales/PHWC-490FB9/2020\|EPI_ISL_706595\|2020 | ···················· | ···················· | ···················· | ···················· | ···················· | ···················· |
| W | 1250 | hCoV-19/Wales/PHWC-490FC8/2020\|EPI_ISL_706596\|2020 | ···················· | ···················· | ···················· | ···················· | ···················· | ···················· |
| W | 1251 | hCoV-19/Wales/PHWC-490FD7/2020\|EPI_ISL_706597\|2020 | ···················· | ···················· | ···················· | ···················· | ···················· | ···················· |
| W | 1252 | hCoV-19/Wales/PHWC-490FE6/2020\|EPI_ISL_706598\|2020 | ···················· | ···················· | ···················· | ···················· | ···················· | ···················· |
| W | 1253 | hCoV-19/Wales/PHWC-49101F/2020\|EPI_ISL_706599\|2020 | ···················· | ······A············· | ···················· | ···················· | ···················· | ···················· |
| W | 1254 | hCoV-19/USA/MI-UM-10036787921/2020\|EPI_ISL_707860\| | ···················· | ···················· | ···················· | ···················· | ···················· | ···················· |
| W | 1259 | hCoV-19/USA/MI-UM-10036789788/2020\|EPI_ISL_707865\| | ···················· | ···················· | ···················· | ···················· | ···················· | ···················· |
| W | 1260 | hCoV-19/USA/MI-UM-10036782417/2020\|EPI_ISL_707866\| | ···················· | ···················· | ···················· | ···················· | ···················· | ···················· |
| W | 1261 | hCoV-19/USA/MI-UM-10036775247/2020\|EPI_ISL_707867\| | ···················· | ···················· | ···················· | ···················· | ···················· | ···················· |
| W | 1263 | hCoV-19/USA/MI-UM-10036794049/2020\|EPI_ISL_707869\| | ···················· | ···················· | ···················· | ···················· | ···················· | ···················· |
| W | 1264 | hCoV-19/USA/MI-UM-10036790151/2020\|EPI_ISL_707870\| | ···················· | ·A·················· | ···················· | ···················· | ···················· | ···················· |
| W | 1265 | hCoV-19/USA/MI-UM-10036789875/2020\|EPI_ISL_707871\| | ···················· | ·A·················· | ···················· | ···················· | ···················· | ···················· |
| W | 1266 | hCoV-19/USA/MI-UM-10036774822/2020\|EPI_ISL_707872\| | ···················· | ···················· | ···················· | ···················· | ···················· | ···················· |
| W | 1267 | hCoV-19/USA/MI-UM-10036782210/2020\|EPI_ISL_707873\| | ···················· | ···················· | ···················· | ···················· | ···················· | ···················· |
| W | 1268 | hCoV-19/USA/MI-UM-10036782077/2020\|EPI_ISL_707874\| | ···················· | ···················· | ···················· | ···················· | ···················· | ···················· |
| W | 1270 | hCoV-19/USA/MI-UM-10036782134/2020\|EPI_ISL_707876\| | ···················· | ·············A······ | ···················· | ···················· | ···················· | ···················· |
| W | 1271 | hCoV-19/USA/MI-UM-10036795528/2020\|EPI_ISL_707877\| | ···················· | ·············A······ | ···················· | ···················· | ···················· | ···················· |
| W | 1272 | hCoV-19/USA/MI-UM-10036785789/2020\|EPI_ISL_707878\| | ···················· | ·············A······ | ···················· | ···················· | ····A··············· | ···················· |
| W | 1273 | hCoV-19/USA/MI-UM-10036784725/2020\|EPI_ISL_707879\| | ···················· | ···················· | ···················· | ···················· | ···················· | ···················· |
| W | 1274 | hCoV-19/Australia/NSW2713/2020\|EPI_ISL_707891\|2020 | ···················· | ···················· | ···················· | ···················· | ···················· | ···················· |
| W | 1275 | hCoV-19/Australia/NSW2714/2020\|EPI_ISL_707892\|2020 | ···················· | ···················· | ···················· | ···················· | ···················· | ···················· |
| W | 1276 | hCoV-19/Australia/NSW2946/2020\|EPI_ISL_707894\|2020 | ···················· | ·A·················· | ···················· | ···················· | ···················· | ···A················ |
| W | 1277 | hCoV-19/Australia/NSW2964/2020\|EPI_ISL_707896\|2020 | ···················· | ·A·················· | ···················· | ···················· | ···················· | ···A················ |
| W | 1278 | hCoV-19/Australia/NSW2967/2020\|EPI_ISL_707897\|2020 | ···················· | ···················· | ···················· | ···················· | ···················· | ···················· |
| W | 1279 | hCoV-19/Australia/NSW2971/2020\|EPI_ISL_707898\|2020 | ···················· | ···················· | ···················· | ···················· | ···················· | ···················· |
| W | 1280 | hCoV-19/Australia/NSW2972/2020\|EPI_ISL_707899\|2020 | ···················· | ·A·················· | ···················· | ···················· | ···················· | ···A················ |
| W | 1281 | hCoV-19/USA/AZ-TG664524/2020\|EPI_ISL_696141\|2020-0 | ···················· | ···················· | ···················· | ···················· | ···················· | ···················· |
| W | 1282 | hCoV-19/USA/AZ-TG664525/2020\|EPI_ISL_696142\|2020-0 | ···················· | ···················· | ···················· | ···················· | ···················· | ···················· |
| W | 1283 | hCoV-19/USA/AZ-TG664526/2020\|EPI_ISL_696143\|2020-0 | ···················· | ···················· | ···················· | ···················· | ···················· | ···················· |
| W | 1284 | hCoV-19/USA/AZ-TG664527/2020\|EPI_ISL_696144\|2020-0 | ···················· | ···················· | ···················· | ···················· | ···················· | ···················· |
| W | 1285 | hCoV-19/USA/AZ-TG664528/2020\|EPI_ISL_696145\|2020-0 | ···················· | ···················· | ···················· | ···················· | ···················· | ···················· |
| W | 1286 | hCoV-19/USA/AZ-TG664532/2020\|EPI_ISL_696149\|2020-0 | ···················· | ···················· | ···················· | ···················· | ···················· | ···················· |
| W | 1287 | hCoV-19/USA/AZ-TG664534/2020\|EPI_ISL_696151\|2020-0 | ···················· | ···················· | ···················· | ···················· | ···················· | ···················· |
| W | 1288 | hCoV-19/USA/AZ-TG664539/2020\|EPI_ISL_696156\|2020-0 | ···················· | ···················· | ···················· | ···················· | ···················· | ···················· |
| W | 1289 | hCoV-19/USA/AZ-TG664541/2020\|EPI_ISL_696158\|2020-0 | ···················· | ···················· | ···················· | ···················· | ···················· | ···················· |
| W | 1290 | hCoV-19/USA/AZ-TG664542/2020\|EPI_ISL_696159\|2020-0 | ···················· | ···················· | ···················· | ···················· | ···················· | ···················· |
| W | 1291 | hCoV-19/USA/AZ-TG664543/2020\|EPI_ISL_696160\|2020-0 | ···················· | ···················· | ···················· | ···················· | ···················· | ···················· |
| W | 1292 | hCoV-19/USA/AZ-TG664549/2020\|EPI_ISL_696166\|2020-0 | ···················· | ···················· | ···················· | ···················· | ···················· | ···················· |
| W | 1293 | hCoV-19/USA/AZ-TG664551/2020\|EPI_ISL_696167\|2020-0 | ···················· | ···················· | ···················· | ···················· | ···················· | ···················· |
| W | 1294 | hCoV-19/USA/AZ-TG664552/2020\|EPI_ISL_696168\|2020-0 | ···················· | ···················· | ···················· | ···················· | ···················· | ···················· |
| W | 1295 | hCoV-19/USA/AZ-TG664553/2020\|EPI_ISL_696169\|2020-0 | ···················· | ···················· | ···················· | ···················· | ···················· | ···················· |
| W | 1296 | hCoV-19/USA/AZ-TG664554/2020\|EPI_ISL_696170\|2020-0 | ···················· | ···················· | ···················· | ···················· | ···················· | ···················· |
| W | 1297 | hCoV-19/USA/AZ-TG664555/2020\|EPI_ISL_696171\|2020-0 | ···················· | ···················· | ···················· | ···················· | ···················· | ···················· |
| W | 1298 | hCoV-19/USA/AZ-TG664558/2020\|EPI_ISL_696173\|2020-0 | ···················· | ···················· | ···················· | ···················· | ···················· | ···················· |
| W | 1299 | hCoV-19/USA/AZ-TG664561/2020\|EPI_ISL_696176\|2020-0 | ···················· | ···················· | ···················· | ···················· | ···················· | ···················· |
| W | 1300 | hCoV-19/USA/AZ-TG666090/2020\|EPI_ISL_696181\|2020-0 | ···················· | ···················· | ···················· | ···················· | ···················· | ···················· |
| W | 1301 | hCoV-19/USA/AZ-TG666095/2020\|EPI_ISL_696185\|2020-0 | ···················· | ···················· | ···················· | ···················· | ···················· | ···················· |
| W | 1302 | hCoV-19/USA/AZ-TG666099/2020\|EPI_ISL_696189\|2020-0 | ···················· | ···················· | ···················· | ···················· | ···················· | ···················· |
| W | 1303 | hCoV-19/USA/AZ-TG666103/2020\|EPI_ISL_696193\|2020-0 | ···················· | ···················· | ···················· | ···················· | ···················· | ···················· |
| W | 1304 | hCoV-19/USA/AZ-TG666104/2020\|EPI_ISL_696194\|2020-0 | ···················· | ·A·················· | ···················· | ···················· | ···················· | ···················· |
| W | 1305 | hCoV-19/USA/AZ-TG666105/2020\|EPI_ISL_696195\|2020-0 | ···················· | ···················· | ···················· | ···················· | ···················· | ···················· |
| W | 1306 | hCoV-19/United Arab Emirates/0550/2020\|EPI_ISL_698 | ···················· | ···················· | ···················· | ···················· | ···················· | ···················· |
| W | 1307 | hCoV-19/United Arab Emirates/0551/2020\|EPI_ISL_698 | ···················· | ···················· | ···················· | ···················· | ···················· | ···················· |
| W | 1308 | hCoV-19/United Arab Emirates/0552/2020\|EPI_ISL_698 | ···················· | ···················· | ···················· | ···················· | ···················· | ···················· |
| W | 1309 | hCoV-19/United Arab Emirates/0553/2020\|EPI_ISL_698 | ···················· | ···················· | ···················· | ···················· | ···················· | ···················· |
| W | 1310 | hCoV-19/United Arab Emirates/0554/2020\|EPI_ISL_698 | ···················· | ···················· | ···················· | ···················· | ···················· | ···················· |
| W | 1311 | hCoV-19/United Arab Emirates/0555/2020\|EPI_ISL_698 | ···················· | ···················· | ···················· | ···················· | ···················· | ···················· |
| W | 1312 | hCoV-19/United Arab Emirates/0556/2020\|EPI_ISL_698 | ···················· | ···················· | ···················· | ···················· | ···················· | ···················· |
| W | 1313 | hCoV-19/United Arab Emirates/0557/2020\|EPI_ISL_698 | ···················· | ···················· | ···················· | ···················· | ···················· | ···················· |
| W | 1314 | hCoV-19/United Arab Emirates/0558/2020\|EPI_ISL_698 | ···················· | ···················· | ···················· | ···················· | ···················· | ···················· |
| W | 1315 | hCoV-19/United Arab Emirates/0559/2020\|EPI_ISL_698 | ···················· | ···················· | ···················· | ···················· | ···················· | ···················· |
| W | 1316 | hCoV-19/United Arab Emirates/0561/2020\|EPI_ISL_698 | ···················· | ···················· | ···················· | ···················· | ···················· | ···················· |
| W | 1317 | hCoV-19/United Arab Emirates/0643/2020\|EPI_ISL_698 | ···················· | ···················· | ···················· | ···················· | ···················· | ···················· |
| W | 1318 | hCoV-19/France/ARA-SC141/2020\|EPI_ISL_700405\|2020- | ···················· | ·A·················· | ···················· | ···················· | ···················· | ···················· |
| W | 1319 | hCoV-19/France/ARA-SC142/2020\|EPI_ISL_700406\|2020- | ···················· | ·A·················· | ···················· | ···················· | ···················· | ···················· |

| | | | | | | | | |
|---|---|---|---|---|---|---|---|---|
| W | 1321 hCoV-19/South Africa/NHLS-UCT-GS-1067/2020\|EPI_ISL | ···················· | ···················· | ···················· | ···················· | ···················· | ···················· | |
| W | 1325 hCoV-19/South Africa/NHLS-UCT-GS-1358/2020\|EPI_ISL | ···················· | ···················· | ···················· | ···················· | ···················· | ···················· | |
| W | 1331 hCoV-19/South Africa/NHLS-UCT-GS-0660/2020\|EPI_ISL | ···················· | ···················· | ···················· | ···················· | ···················· | ···················· | |
| W | 1333 hCoV-19/South Africa/NHLS-UCT-GS-1350/2020\|EPI_ISL | ···················· | ···················· | ···················· | ···················· | ···················· | ···················· | |
| W | 1339 hCoV-19/South Africa/NHLS-UCT-GS-1576/2020\|EPI_ISL | ···················· | ·A·················· | ···················· | ···················· | ···················· | ···················· | |
| W | 1340 hCoV-19/South Africa/NHLS-UCT-GS-1525/2020\|EPI_ISL | ···················· | ···················· | ···················· | ···················· | ···················· | ···················· | |
| W | 1341 hCoV-19/South Africa/NHLS-UCT-GS-1354/2020\|EPI_ISL | ···················· | ·A·················· | ···················· | ···················· | ···················· | ···················· | |
| W | 1342 hCoV-19/South Africa/NHLS-UCT-GP-5156/2020\|EPI_ISL | ···················· | ·A·················· | ···················· | ···················· | ···················· | ···················· | |
| W | 1345 hCoV-19/South Africa/NHLS-UCT-GP-5386/2020\|EPI_ISL | ···················· | ·A·················· | ···················· | ···················· | ···················· | ···················· | |
| W | 1350 hCoV-19/South Africa/NHLS-UCT-GP-5664/2020\|EPI_ISL | ···················· | ·A·················· | ···················· | ···················· | ···················· | ···················· | |
| W | 1351 hCoV-19/South Africa/NHLS-UCT-GS-3258/2020\|EPI_ISL | ···················· | ·A·················· | ···················· | ···················· | ···················· | ···················· | |
| W | 1353 hCoV-19/South Africa/NHLS-UCT-GS-1532/2020\|EPI_ISL | ···················· | ···················· | ···················· | ···················· | ···················· | ···················· | |
| W | 1354 hCoV-19/South Africa/NHLS-UCT-GS-2214/2020\|EPI_ISL | ···················· | ·A·················· | ···················· | ···················· | ···················· | ···················· | |
| W | 1356 hCoV-19/South Africa/NHLS-UCT-GS-1078/2020\|EPI_ISL | ···················· | ···················· | ···················· | ···················· | ···················· | ···················· | |
| W | 1359 hCoV-19/South Africa/NHLS-UCT-GP-5256/2020\|EPI_ISL | ···················· | ···················· | ···················· | ···················· | ···················· | ···················· | |
| W | 1360 hCoV-19/South Africa/NHLS-UCT-GP-5422/2020\|EPI_ISL | ···················· | ···················· | ···················· | ···················· | ···················· | ···················· | |
| W | 1361 hCoV-19/South Africa/NHLS-UCT-GP-5818/2020\|EPI_ISL | ···················· | ···················· | ···················· | ···················· | ·NNN················ | ···················· | 1 |
| W | 1362 hCoV-19/South Africa/NHLS-UCT-GS-1368/2020\|EPI_ISL | ···················· | ·A·················· | ···················· | ···················· | ···················· | ···················· | |
| W | 1364 hCoV-19/South Africa/NHLS-UCT-GS-0690/2020\|EPI_ISL | ···················· | ·A·················· | ···················· | ···················· | ···················· | ···················· | |
| W | 1365 hCoV-19/South Africa/NHLS-UCT-GP-5912/2020\|EPI_ISL | ···················· | ···················· | ···················· | ···················· | ···················· | ···················· | |
| W | 1367 hCoV-19/South Africa/NHLS-UCT-GS-2232/2020\|EPI_ISL | ···················· | ···················· | ···················· | ···················· | ···················· | ···················· | |
| W | 1368 hCoV-19/South Africa/NHLS-UCT-GS-2452/2020\|EPI_ISL | ···················· | ·A·················· | ···················· | ···················· | ···················· | ···················· | |
| W | 1372 hCoV-19/South Africa/NHLS-UCT-GS-0675/2020\|EPI_ISL | ···················· | ···················· | ···················· | ···················· | ···················· | ···················· | |
| W | 1374 hCoV-19/South Africa/NHLS-UCT-GP-5241/2020\|EPI_ISL | ···················· | ·A·················· | ···················· | ···················· | ···················· | ···················· | |
| W | 1376 hCoV-19/South Africa/NHLS-UCT-GS-1259/2020\|EPI_ISL | ···················· | ···················· | ···················· | ···················· | ···················· | ···················· | |
| W | 1377 hCoV-19/South Africa/NHLS-UCT-GS-2116/2020\|EPI_ISL | ···················· | ·A·················· | ···················· | ···················· | ···················· | ···················· | |
| W | 1379 hCoV-19/South Africa/NHLS-UCT-GS-3097/2020\|EPI_ISL | ···················· | ···················· | ···················· | ···················· | ···················· | ···················· | |
| W | 1381 hCoV-19/South Africa/NHLS-UCT-GS-1648/2020\|EPI_ISL | ···················· | ·A·················· | ···················· | ···················· | ···················· | ···················· | |
| W | 1382 hCoV-19/South Africa/NHLS-UCT-GP-5388/2020\|EPI_ISL | ···················· | ·A·················· | ···················· | ···················· | ···················· | ···················· | |
| W | 1383 hCoV-19/South Africa/NHLS-UCT-GS-2217/2020\|EPI_ISL | ···················· | ···················· | ···················· | ···················· | ···················· | ···················· | |
| W | 1385 hCoV-19/South Africa/NHLS-UCT-GS-1026/2020\|EPI_ISL | ···················· | ·A·················· | ···················· | ···················· | ···················· | ···················· | |
| W | 1387 hCoV-19/South Africa/NHLS-UCT-GP-5742/2020\|EPI_ISL | ···················· | ···················· | ···················· | ···················· | ···················· | ···················· | |
| W | 1388 hCoV-19/England/ALDP-BBB467/2020\|EPI_ISL_701758\|20 | ···················· | ···················· | ···················· | ···················· | ···················· | ···A················ | |
| W | 1389 hCoV-19/England/ALDP-BB3C76/2020\|EPI_ISL_701759\|20 | ···················· | ···················· | ···················· | ···················· | ······T············· | ···················· | |
| W | 1390 hCoV-19/England/ALDP-BBA42C/2020\|EPI_ISL_701760\|20 | ···················· | ···················· | ···················· | ···················· | ···················· | ···················· | |
| W | 1391 hCoV-19/England/ALDP-BB5175/2020\|EPI_ISL_701761\|20 | ···················· | ···················· | ···················· | ···················· | ···················· | ···················· | |
| W | 1392 hCoV-19/England/ALDP-BB3599/2020\|EPI_ISL_701762\|20 | ···················· | ···················· | ···················· | ···················· | ···················· | ···················· | |
| W | 1393 hCoV-19/England/ALDP-BBB2D6/2020\|EPI_ISL_701763\|20 | ···················· | ···················· | ···················· | ···················· | ···················· | ···················· | |
| W | 1394 hCoV-19/England/ALDP-BBA170/2020\|EPI_ISL_701764\|20 | ···················· | ···················· | ···················· | ···················· | ···················· | ···················· | |
| W | 1395 hCoV-19/England/ALDP-BBB42B/2020\|EPI_ISL_701766\|20 | ···················· | ···················· | ···················· | ···················· | ···················· | ···················· | |
| W | 1396 hCoV-19/England/ALDP-BBA8B7/2020\|EPI_ISL_701768\|20 | ···················· | ·A·················· | ···················· | ···················· | ···················· | ···A················ | |
| W | 1397 hCoV-19/England/ALDP-BBAC7F/2020\|EPI_ISL_701769\|20 | ···················· | ···················· | ···················· | ···················· | ···················· | ···················· | |
| W | 1398 hCoV-19/England/ALDP-BB5412/2020\|EPI_ISL_701770\|20 | ···················· | ···················· | ···················· | ···················· | ···················· | ···················· | |
| W | 1399 hCoV-19/England/ALDP-BB9F59/2020\|EPI_ISL_701771\|20 | ···················· | ···················· | ···················· | ···················· | ···················· | ···················· | |
| W | 1400 hCoV-19/England/ALDP-BB9E5C/2020\|EPI_ISL_701773\|20 | ···················· | ···················· | ···················· | ···················· | ···················· | ···················· | |
| W | 1401 hCoV-19/England/ALDP-BB3863/2020\|EPI_ISL_701774\|20 | ···················· | ···················· | ···················· | ···················· | ···················· | ···················· | |
| W | 1402 hCoV-19/England/ALDP-BBB133/2020\|EPI_ISL_701777\|20 | ···················· | ·········A·········· | ···················· | ···················· | ···················· | ···················· | |
| W | 1403 hCoV-19/England/ALDP-BBAFD0/2020\|EPI_ISL_701778\|20 | ···················· | ···················· | ···················· | ···················· | ···················· | ···················· | |
| W | 1404 hCoV-19/England/ALDP-BBB34C/2020\|EPI_ISL_701779\|20 | ···················· | ···················· | ···················· | ···················· | ···················· | ···················· | |
| W | 1405 hCoV-19/England/ALDP-BB38AF/2020\|EPI_ISL_701780\|20 | ···················· | ···················· | ···················· | ···················· | ···················· | ···················· | |
| W | 1406 hCoV-19/Scotland/ALDP-BB3766/2020\|EPI_ISL_701781\|2 | ···················· | ···················· | ···················· | ···················· | ···················· | ···················· | |
| W | 1407 hCoV-19/England/ALDP-BB3881/2020\|EPI_ISL_701783\|20 | ···················· | ···················· | ···················· | ···················· | ···················· | ···················· | |
| W | 1408 hCoV-19/England/ALDP-BBB230/2020\|EPI_ISL_701785\|20 | ···················· | ···················· | ···················· | ···················· | ···················· | ···················· | |
| W | 1409 hCoV-19/England/ALDP-BBA8F3/2020\|EPI_ISL_701786\|20 | ···················· | ···················· | ···················· | ···················· | ···················· | ···················· | |
| W | 1410 hCoV-19/England/ALDP-BBB4C1/2020\|EPI_ISL_701787\|20 | ···················· | ···················· | ···················· | ···················· | ···················· | ···················· | |
| W | 1411 hCoV-19/England/ALDP-BB3C0D/2020\|EPI_ISL_701788\|20 | ···················· | ···················· | ···················· | ···················· | ···················· | ···················· | |
| W | 1412 hCoV-19/England/ALDP-BB3775/2020\|EPI_ISL_701789\|20 | ···················· | ···················· | ···················· | ···················· | ···················· | ···················· | |
| W | 1413 hCoV-19/England/ALDP-BBB43A/2020\|EPI_ISL_701790\|20 | ···················· | ···················· | ···················· | ···················· | ···················· | ···················· | |
| W | 1414 hCoV-19/England/ALDP-BB5254/2020\|EPI_ISL_701791\|20 | ···················· | ···················· | ···················· | ···················· | ···················· | ···················· | |
| W | 1415 hCoV-19/England/ALDP-BB38CD/2020\|EPI_ISL_701793\|20 | ···················· | ···················· | ···················· | ···················· | ···················· | ···················· | |
| W | 1416 hCoV-19/England/ALDP-BB9E2F/2020\|EPI_ISL_701794\|20 | ···················· | ···················· | ···················· | ···················· | ···················· | ···················· | |
| W | 1417 hCoV-19/England/ALDP-BB3F8C/2020\|EPI_ISL_701795\|20 | ···················· | ···················· | ···················· | ···················· | ···················· | ···················· | |
| W | 1418 hCoV-19/England/ALDP-BB9F4A/2020\|EPI_ISL_701796\|20 | ···················· | ···················· | ···················· | ···················· | ···················· | ···················· | |
| W | 1419 hCoV-19/England/ALDP-BBAF67/2020\|EPI_ISL_701797\|20 | ···················· | ···················· | ···················· | ···················· | ···················· | ···················· | |
| W | 1420 hCoV-19/England/ALDP-BB3D64/2020\|EPI_ISL_701798\|20 | ···················· | ···················· | ···················· | ···················· | ···················· | ···················· | |
| W | 1421 hCoV-19/England/ALDP-BB5227/2020\|EPI_ISL_701799\|20 | ···················· | ···················· | ···················· | ···················· | ···················· | ···················· | |
| W | 1422 hCoV-19/England/QEUH-BC3675/2020\|EPI_ISL_704010\|20 | ···················· | ·A·················· | ···················· | ···················· | ···················· | ···A················ | |
| W | 1423 hCoV-19/England/QEUH-BC3AE2/2020\|EPI_ISL_704013\|20 | ···················· | ···················· | ···················· | ···················· | ···················· | ···················· | |
| W | 1424 hCoV-19/England/QEUH-BCB84A/2020\|EPI_ISL_704015\|20 | ···················· | ···················· | ···················· | ···················· | ···················· | ···················· | |
| W | 1425 hCoV-19/England/QEUH-BC3754/2020\|EPI_ISL_704016\|20 | ···················· | ···················· | ···················· | ···················· | ···················· | ···················· | |
| W | 1426 hCoV-19/England/QEUH-BCBC5D/2020\|EPI_ISL_704019\|20 | ···················· | ·A·················· | ···················· | ···················· | ···················· | ···A················ | |
| W | 1428 hCoV-19/Wales/PHWC-49103D/2020\|EPI_ISL_706601\|2020 | ···················· | ···················· | ···················· | ···················· | ···················· | ···················· | |
| W | 1429 hCoV-19/Wales/PHWC-49104C/2020\|EPI_ISL_706602\|2020 | ···················· | ···················· | ···················· | ···················· | ···················· | ···················· | |
| W | 1430 hCoV-19/Wales/PHWC-49105B/2020\|EPI_ISL_706603\|2020 | ···················· | ···················· | ···················· | ···················· | NNN················· | ···················· | 1 |
| W | 1433 hCoV-19/Wales/PHWC-491097/2020\|EPI_ISL_706606\|2020 | ···················· | ···················· | ···················· | ···················· | ···················· | ···················· | |
| W | 1436 hCoV-19/Wales/PHWC-4910C4/2020\|EPI_ISL_706609\|2020 | ···················· | ···················· | ···················· | ···················· | ···················· | ···················· | |
| W | 1438 hCoV-19/Wales/PHWC-4910E2/2020\|EPI_ISL_706611\|2020 | ···················· | ···················· | ···················· | ···················· | ···················· | ···················· | |
| W | 1439 hCoV-19/Wales/PHWC-4910F1/2020\|EPI_ISL_706612\|2020 | ···················· | ···················· | ···················· | ···················· | ···················· | ···················· | |
| W | 1440 hCoV-19/Wales/PHWC-49111C/2020\|EPI_ISL_706613\|2020 | ···················· | ···················· | ···················· | ···················· | ···················· | ···················· | |
| W | 1441 hCoV-19/Wales/PHWC-49112B/2020\|EPI_ISL_706614\|2020 | ···················· | ···················· | ···················· | ···················· | ···················· | ···················· | |

| | | | | | | | | |
|---|---|---|---|---|---|---|---|---|
| W | 1442 hCoV-19/Wales/PHWC-49113A/2020\|EPI_ISL_706615\|2020 | ···················· | ···················· | ···················· | ···················· | ···················· | ···················· | |
| W | 1444 hCoV-19/Wales/PHWC-491158/2020\|EPI_ISL_706617\|2020 | ···················· | ···················· | ···················· | ···················· | ···················· | ···················· | |
| W | 1445 hCoV-19/Wales/PHWC-491167/2020\|EPI_ISL_706618\|2020 | ···················· | ···················· | ···················· | ···················· | ···················· | ···················· | |
| W | 1446 hCoV-19/Wales/PHWC-491176/2020\|EPI_ISL_706619\|2020 | ···················· | ···················· | ···················· | ···················· | ···················· | ···················· | |
| W | 1447 hCoV-19/Wales/PHWC-491194/2020\|EPI_ISL_706620\|2020 | ···················· | ···················· | ···················· | ···················· | ···················· | ···················· | |
| W | 1449 hCoV-19/Wales/PHWC-4911B2/2020\|EPI_ISL_706622\|2020 | ···················· | ···················· | ···················· | ···················· | ···················· | ···················· | |
| W | 1450 hCoV-19/Wales/PHWC-4911C1/2020\|EPI_ISL_706623\|2020 | ···················· | ···················· | ···················· | ···················· | ···················· | ···················· | |
| W | 1452 hCoV-19/Wales/PHWC-4911FE/2020\|EPI_ISL_706625\|2020 | ···················· | ···················· | ···················· | ···················· | ···················· | ···················· | |
| W | 1453 hCoV-19/Wales/PHWC-49120A/2020\|EPI_ISL_706626\|2020 | ···················· | ···················· | ···················· | ···················· | ····T··············· | ···················· | |
| W | 1454 hCoV-19/Wales/PHWC-491219/2020\|EPI_ISL_706627\|2020 | ···················· | ···················· | ···················· | ···················· | ···················· | ···················· | |
| W | 1455 hCoV-19/Wales/PHWC-491228/2020\|EPI_ISL_706628\|2020 | ···················· | ···················· | ···················· | ···················· | ···················· | ···················· | |
| W | 1456 hCoV-19/Wales/PHWC-491237/2020\|EPI_ISL_706629\|2020 | ···················· | ···················· | ···················· | ···················· | ···················· | ···················· | |
| W | 1457 hCoV-19/Wales/PHWC-491246/2020\|EPI_ISL_706630\|2020 | ···················· | ···················· | ···················· | ···················· | ···················· | ···················· | |
| W | 1458 hCoV-19/Wales/PHWC-491255/2020\|EPI_ISL_706631\|2020 | ···················· | ···················· | ···················· | ···················· | ···················· | ···················· | |
| W | 1459 hCoV-19/Wales/PHWC-491264/2020\|EPI_ISL_706632\|2020 | ···················· | ···················· | ···················· | ···················· | ···················· | ···················· | |
| W | 1460 hCoV-19/Wales/PHWC-491273/2020\|EPI_ISL_706633\|2020 | ···················· | ···················· | ···················· | ···················· | ···················· | ···················· | |
| W | 1463 hCoV-19/Wales/PHWC-4912BF/2020\|EPI_ISL_706636\|2020 | ···················· | ···················· | ···················· | ···················· | ···················· | ···················· | |
| W | 1464 hCoV-19/Wales/PHWC-4912CE/2020\|EPI_ISL_706637\|2020 | ···················· | ···················· | ···················· | ···················· | ···················· | ···················· | |
| W | 1465 hCoV-19/Wales/PHWC-4912DD/2020\|EPI_ISL_706638\|2020 | ···················· | ···················· | ···················· | ···················· | ···················· | ···················· | |
| W | 1466 hCoV-19/Wales/PHWC-4912EC/2020\|EPI_ISL_706639\|2020 | ···················· | ···················· | ···················· | ···················· | ···················· | ···················· | |
| W | 1467 hCoV-19/Wales/PHWC-4912FB/2020\|EPI_ISL_706640\|2020 | ···················· | ···················· | ···················· | ···················· | ···················· | ···················· | |
| W | 1468 hCoV-19/Wales/PHWC-491307/2020\|EPI_ISL_706641\|2020 | ···················· | ···················· | ···················· | ···················· | ···················· | ···················· | |
| W | 1470 hCoV-19/Wales/PHWC-491325/2020\|EPI_ISL_706643\|2020 | ···················· | ···················· | ···················· | ···················· | ···················· | ···················· | |
| W | 1471 hCoV-19/Wales/PHWC-491334/2020\|EPI_ISL_706644\|2020 | ···················· | ···················· | ···················· | ···················· | ···················· | ···················· | |
| W | 1472 hCoV-19/Wales/PHWC-491370/2020\|EPI_ISL_706645\|2020 | ···················· | ···················· | ···················· | ···················· | NNN················· | ····NN·············· | 1 |
| W | 1473 hCoV-19/Wales/PHWC-49138F/2020\|EPI_ISL_706646\|2020 | ···················· | ···················· | ···················· | ···················· | ···················· | ···················· | |
| W | 1474 hCoV-19/Wales/PHWC-49139E/2020\|EPI_ISL_706647\|2020 | ···················· | ···················· | ···················· | ···················· | ···················· | ···················· | |
| W | 1476 hCoV-19/Wales/PHWC-4913DA/2020\|EPI_ISL_706649\|2020 | ···················· | ···················· | ···················· | ···················· | ···················· | ···················· | |
| W | 1482 hCoV-19/Wales/PHWC-491668/2020\|EPI_ISL_706675\|2020 | ···················· | ···················· | ···················· | ···················· | ···················· | ···················· | |
| W | 1483 hCoV-19/Wales/PHWC-491677/2020\|EPI_ISL_706676\|2020 | ···················· | ···················· | ···················· | ···················· | ···················· | ···················· | |
| W | 1484 hCoV-19/Wales/PHWC-491686/2020\|EPI_ISL_706677\|2020 | ···················· | ···················· | ···················· | ···················· | ···················· | ···················· | |
| W | 1485 hCoV-19/Australia/NSW2977/2020\|EPI_ISL_707900\|2020 | ···················· | ···················· | ···················· | ···················· | ···················· | ···················· | |
| W | 1486 hCoV-19/Australia/NSW2979/2020\|EPI_ISL_707901\|2020 | ···················· | ····G··············· | ···················· | ···················· | ···················· | ···················· | |
| W | 1487 hCoV-19/Australia/NSW2985/2020\|EPI_ISL_707902\|2020 | ···················· | ···················· | ···················· | ···················· | ···················· | ···················· | |
| W | 1488 hCoV-19/USA/CA-LACPHL-AE00001/2020\|EPI_ISL_707903\| | ···················· | ···················· | ···················· | ···················· | ···················· | ···················· | |
| W | 1489 hCoV-19/USA/CA-LACPHL-AE00003/2020\|EPI_ISL_707904\| | ···················· | ···················· | ···················· | ···················· | ···················· | ···················· | |
| W | 1490 hCoV-19/USA/CA-LACPHL-AE00004/2020\|EPI_ISL_707905\| | ···················· | ···················· | ···················· | ···················· | ···················· | ···················· | |
| W | 1491 hCoV-19/USA/CA-LACPHL-AE00007/2020\|EPI_ISL_707906\| | ···················· | ···················· | ···················· | ···················· | ···················· | ···················· | |
| W | 1492 hCoV-19/USA/CA-LACPHL-AE00009/2020\|EPI_ISL_707907\| | ···················· | ···················· | ···················· | ···················· | ···················· | ···················· | |
| W | 1493 hCoV-19/USA/CA-LACPHL-AE00011/2020\|EPI_ISL_707908\| | ···················· | ···················· | ···················· | ···················· | ···················· | ···················· | |
| W | 1494 hCoV-19/USA/CA-LACPHL-AE00012/2020\|EPI_ISL_707909\| | ···················· | ···················· | ···················· | ···················· | ···················· | ···················· | |
| W | 1495 hCoV-19/USA/CA-LACPHL-AE00013/2020\|EPI_ISL_707910\| | ···················· | ·A·················· | ···················· | ···················· | ···················· | ···················· | |
| W | 1496 hCoV-19/USA/CA-LACPHL-AE00014/2020\|EPI_ISL_707911\| | ···················· | ·A·················· | ···················· | ···················· | ···················· | ···················· | |
| W | 1497 hCoV-19/USA/CA-LACPHL-AE00015/2020\|EPI_ISL_707912\| | ···················· | ·A·················· | ···················· | ···················· | ···················· | ···················· | |
| W | 1498 hCoV-19/USA/CA-LACPHL-AE00016/2020\|EPI_ISL_707913\| | ···················· | ···················· | ···················· | ···················· | ···················· | ···················· | |
| W | 1499 hCoV-19/USA/CA-LACPHL-AE00017/2020\|EPI_ISL_707914\| | ···················· | ···················· | ···················· | ···················· | ···················· | ···················· | |
| W | 1500 hCoV-19/USA/CA-LACPHL-AE00018/2020\|EPI_ISL_707915\| | ···················· | ···················· | ···················· | ···················· | ···················· | ···················· | |
| W | 1501 hCoV-19/USA/CA-LACPHL-AE00019/2020\|EPI_ISL_707916\| | ···················· | ·A·················· | ···················· | ···················· | ···················· | ···················· | |
| W | 1502 hCoV-19/USA/CA-LACPHL-AE00020/2020\|EPI_ISL_707917\| | ···················· | ···················· | ···················· | ···················· | ···················· | ···················· | |
| W | 1503 hCoV-19/USA/CA-LACPHL-AE00021/2020\|EPI_ISL_707918\| | ···················· | ·A·················· | ···················· | ···················· | ···················· | ···················· | |
| W | 1504 hCoV-19/USA/CA-LACPHL-AE00022/2020\|EPI_ISL_707919\| | ···················· | ···················· | ···················· | ···················· | ···················· | ···················· | |
| W | 1505 hCoV-19/USA/CA-LACPHL-AE00023/2020\|EPI_ISL_707920\| | ···················· | ···················· | ···················· | ···················· | ···················· | ···················· | |
| W | 1506 hCoV-19/USA/CA-LACPHL-AE00024/2020\|EPI_ISL_707921\| | ···················· | ·A·················· | ···················· | ···················· | ···················· | ···················· | |
| W | 1507 hCoV-19/USA/CA-LACPHL-AE00025/2020\|EPI_ISL_707922\| | ···················· | ···················· | ···················· | ···················· | ···················· | ···················· | |
| W | 1508 hCoV-19/USA/CA-LACPHL-AE00026/2020\|EPI_ISL_707923\| | ···········T········ | ···················· | ···················· | ···················· | ···················· | ···················· | |
| W | 1509 hCoV-19/USA/CA-LACPHL-AE00027/2020\|EPI_ISL_707924\| | ···················· | ····T··············· | ···················· | ···················· | ···················· | ···················· | |
| W | 1510 hCoV-19/USA/CA-LACPHL-AE00028/2020\|EPI_ISL_707925\| | ···················· | ·A·················· | ···················· | ···················· | ···················· | ···················· | |
| W | 1511 hCoV-19/USA/CA-LACPHL-AE00031/2020\|EPI_ISL_707926\| | ···················· | ····T··············· | ···················· | ···················· | ···················· | ···················· | |
| W | 1512 hCoV-19/USA/CA-LACPHL-AE00002/2020\|EPI_ISL_707927\| | ···················· | ····T··············· | ···················· | ···················· | ···················· | ···················· | |
| W | 1513 hCoV-19/Japan/FHU-Cor529/2020\|EPI_ISL_707928\|2020- | ···················· | ···················· | ···················· | ···················· | ···················· | ···················· | |
| W | 1514 hCoV-19/USA/CA-LACPHL-AE00005/2020\|EPI_ISL_707929\| | ···················· | ···················· | ···················· | ···················· | ···················· | ···················· | |
| W | 1515 hCoV-19/USA/CA-LACPHL-AE00008/2020\|EPI_ISL_707930\| | ···················· | ···················· | ···················· | ···················· | ···················· | ···················· | |
| W | 1516 hCoV-19/USA/CA-LACPHL-AE00029/2020\|EPI_ISL_707931\| | ···················· | ···················· | ···················· | ···················· | ···················· | ···················· | |
| W | 1517 hCoV-19/USA/CA-LACPHL-AE00030/2020\|EPI_ISL_707932\| | ···················· | ····T··············· | ···················· | ···················· | ···················· | ···················· | |
| W | 1518 hCoV-19/Turkey/DEN-PAU05/2020\|EPI_ISL_707936\|2020- | ···················· | ···················· | ···················· | ···················· | ···················· | ···················· | |
| W | 1519 hCoV-19/Turkey/DEN-PAU07/2020\|EPI_ISL_707937\|2020- | ···················· | ···················· | ···················· | ···················· | ···················· | ···················· | |
| W | 1520 hCoV-19/Turkey/DEN-PAU01/2020\|EPI_ISL_707938\|2020- | ···················· | ···················· | ···················· | ···················· | ···················· | ···················· | |
| W | 1521 hCoV-19/Turkey/DEN-PAU02/2020\|EPI_ISL_707939\|2020- | ···················· | ···················· | ···················· | ···················· | ···················· | ···················· | |
| W | 1522 hCoV-19/Germany/SL-SU-10426887/2020\|EPI_ISL_707943 | ···················· | ···················· | ···················· | ···················· | ···················· | ···················· | |
| W | 1523 hCoV-19/Germany/SL-SU-10427230/2020\|EPI_ISL_707947 | ···················· | ···················· | ···················· | ···················· | ···················· | ···················· | |
| W | 1524 hCoV-19/Germany/SL-SU-10427348/2020\|EPI_ISL_707948 | ···················· | ·············A······ | ···················· | ···················· | ···················· | ···················· | |
| W | 1525 hCoV-19/Germany/SL-SU-10427535/2020\|EPI_ISL_707950 | ···················· | ···················· | ···················· | ···················· | ···················· | ···················· | |
| W | 1526 hCoV-19/Germany/SL-SU-10427552/2020\|EPI_ISL_707951 | ···················· | ···················· | ···················· | ···················· | ···················· | ···················· | |
| W | 1527 hCoV-19/Germany/SL-SU-10428103/2020\|EPI_ISL_707954 | ···················· | ···················· | ···················· | ···················· | ···················· | ···················· | |
| W | 1528 hCoV-19/Germany/SL-SU-10429159/2020\|EPI_ISL_707956 | ···················· | ···················· | ···················· | ···················· | ···················· | ···············A···· | |
| W | 1529 hCoV-19/Germany/SL-SU-10430142/2020\|EPI_ISL_707959 | ···················· | ···················· | ···················· | ···················· | ···················· | ···················· | |
| W | 1530 hCoV-19/Germany/SL-SU-10431265/2020\|EPI_ISL_707964 | ···················· | ···················· | ···················· | ···················· | ···················· | ···················· | |
| W | 1531 hCoV-19/Germany/SL-SU-10431379/2020\|EPI_ISL_707966 | ···················· | ···················· | ···················· | ···················· | ···················· | ···················· | |
| W | 1532 hCoV-19/Germany/SL-SU-10431588/2020\|EPI_ISL_707968 | ···················· | ···················· | ···················· | ···················· | ···················· | ·············NN····· | |
| W | 1534 hCoV-19/Germany/SL-SU-10450152/2020\|EPI_ISL_707971 | ···················· | ···················· | ···················· | ···················· | ···················· | ···················· | |

W 1535 hCoV-19/Germany/SL-SU-10450307/2020|EPI_ISL_707972 .................... .................... ..................... .................... .................... ....................
W 1536 hCoV-19/Germany/SL-SU-10450502/2020|EPI_ISL_707975 .................... .................... ..................... .................... .................... ....................
W 1537 hCoV-19/USA/AZ-TG664470/2020|EPI_ISL_696103|2020-0 .................... .................... ..................... .................... .................... ....................
W 1538 hCoV-19/USA/AZ-TG664477/2020|EPI_ISL_696107|2020-0 .................... .................... ..................... .................... .................... ....................
W 1539 hCoV-19/USA/AZ-TG664480/2020|EPI_ISL_696110|2020-0 .................... .................... ..................... .................... .................... ....................
W 1540 hCoV-19/USA/AZ-TG664489/2020|EPI_ISL_696115|2020-0 .................... .................... ..................... .................... .................... ....................
W 1541 hCoV-19/USA/AZ-TG664490/2020|EPI_ISL_696116|2020-0 .................... .................... ..................... .................... .................... ....................
W 1542 hCoV-19/USA/AZ-TG664491/2020|EPI_ISL_696117|2020-0 .................... .................... ..................... .................... .................... ....................
W 1543 hCoV-19/USA/AZ-TG664493/2020|EPI_ISL_696119|2020-0 .................... .................... ..................... .................... .................... ....................
W 1544 hCoV-19/USA/AZ-TG664495/2020|EPI_ISL_696120|2020-0 .................... .................... ..................... .................... .................... ....................
W 1545 hCoV-19/USA/AZ-TG664498/2020|EPI_ISL_696123|2020-0 .................... .................... ..................... .................... .................... ....................
W 1546 hCoV-19/USA/AZ-TG664501/2020|EPI_ISL_696125|2020-0 .................... .................... ..................... .................... .................... ....................
W 1547 hCoV-19/USA/AZ-TG664502/2020|EPI_ISL_696126|2020-0 .................... .................... ..................... .................... .................... ....................
W 1548 hCoV-19/USA/AZ-TG664504/2020|EPI_ISL_696128|2020-0 .................... .................... ..................... .................... .................... ....................
W 1549 hCoV-19/USA/AZ-TG664511/2020|EPI_ISL_696133|2020-0 .................... .................... ..................... .................... .................... ....................
W 1550 hCoV-19/USA/AZ-TG664512/2020|EPI_ISL_696134|2020-0 .................... .................... ..................... .................... .................... ....................
W 1551 hCoV-19/United Arab Emirates/0562/2020|EPI_ISL_698 .................... .................... ..................... .................... .................... ....................
W 1552 hCoV-19/United Arab Emirates/0563/2020|EPI_ISL_698 .................... .................... ..................... .................... .................... ....................
W 1553 hCoV-19/United Arab Emirates/0564/2020|EPI_ISL_698 .................... .................... ..................... .................... .................... ....................
W 1554 hCoV-19/United Arab Emirates/0566/2020|EPI_ISL_698 .................... .................... ..................... .................... .................... ....................
W 1555 hCoV-19/United Arab Emirates/0567/2020|EPI_ISL_698 .................... .................... ..................... .................... .................... ....................
W 1556 hCoV-19/United Arab Emirates/0568/2020|EPI_ISL_698 .................... .................... ..................... .................... .................... ....................
W 1557 hCoV-19/United Arab Emirates/0569/2020|EPI_ISL_698 .................... .................... ..................... .................... .................... ....................
W 1558 hCoV-19/United Arab Emirates/0570/2020|EPI_ISL_698 .................... .................... ..................... .................... .................... ....................
W 1559 hCoV-19/United Arab Emirates/0571/2020|EPI_ISL_698 .................... .................... ..................... .................... .................... ....................
W 1560 hCoV-19/United Arab Emirates/0573/2020|EPI_ISL_698 .................... .................... ..................... .................... .................... ....................
W 1561 hCoV-19/United Arab Emirates/0574/2020|EPI_ISL_698 .................... .................... ..................... .................... .................... ....................
W 1562 hCoV-19/United Arab Emirates/0575/2020|EPI_ISL_698 .................... .................... ..................... .................... .................... ....................
W 1563 hCoV-19/United Arab Emirates/0576/2020|EPI_ISL_698 .................... .................... ..................... .................... .................... ....................
W 1564 hCoV-19/United Arab Emirates/0577/2020|EPI_ISL_698 .................... .................... ..................... .................... .................... ....................
W 1565 hCoV-19/United Arab Emirates/0578/2020|EPI_ISL_698 .................... .................... ..................... .................... .................... ....................
W 1566 hCoV-19/United Arab Emirates/0579/2020|EPI_ISL_698 .................... .................... ..................... .................... .................... ....................
W 1567 hCoV-19/United Arab Emirates/0580/2020|EPI_ISL_698 .................... .................... ..................... .................... .................... ....................
W 1568 hCoV-19/United Arab Emirates/0581/2020|EPI_ISL_698 .................... .................... ..................... .................... .................... ....................
W 1569 hCoV-19/United Arab Emirates/0582/2020|EPI_ISL_698 .................... .................... ..................... .................... .................... ....................
W 1570 hCoV-19/United Arab Emirates/0583/2020|EPI_ISL_698 .................... .................... ..................... .................... .................... ....................
W 1571 hCoV-19/United Arab Emirates/0584/2020|EPI_ISL_698 .................... .................... ..................... .................... .................... ....................
W 1572 hCoV-19/United Arab Emirates/0585/2020|EPI_ISL_698 .................... .................... ..................... .................... .................... ....................
W 1573 hCoV-19/United Arab Emirates/0586/2020|EPI_ISL_698 .................... .................... ..................... .................... .................... ....................
W 1574 hCoV-19/United Arab Emirates/0587/2020|EPI_ISL_698 .................... .................... ..................... .................... .................... ....................
W 1575 hCoV-19/United Arab Emirates/0588/2020|EPI_ISL_698 .................... .................... ..................... .................... .................... ....................
W 1576 hCoV-19/United Arab Emirates/0589/2020|EPI_ISL_698 .................... .................... ..................... .................... .................... ....................
W 1577 hCoV-19/United Arab Emirates/0590/2020|EPI_ISL_698 .................... .................... ..................... .................... .................... ....................
W 1578 hCoV-19/United Arab Emirates/0591/2020|EPI_ISL_698 .................... .................... ..................... .................... .................... ....................
W 1579 hCoV-19/United Arab Emirates/0592/2020|EPI_ISL_698 .................... .................... ..................... .................... .................... ....................
W 1580 hCoV-19/United Arab Emirates/0593/2020|EPI_ISL_698 .................... .................... ..................... .................... .................... ....................
W 1581 hCoV-19/United Arab Emirates/0594/2020|EPI_ISL_698 .................... .................... ..................... .................... .................... ....................
W 1582 hCoV-19/United Arab Emirates/0595/2020|EPI_ISL_698 .................... .................... ..................... .................... .................... ....................
W 1583 hCoV-19/United Arab Emirates/0596/2020|EPI_ISL_698 .................... .................... ..................... .................... .................... ....................
W 1584 hCoV-19/United Arab Emirates/0597/2020|EPI_ISL_698 .................... .................... ..................... .................... .................... ....................
W 1585 hCoV-19/United Arab Emirates/0598/2020|EPI_ISL_698 .................... .................... ..................... .................... .................... ....................
W 1586 hCoV-19/United Arab Emirates/0599/2020|EPI_ISL_698 .................... .................... ..................... .................... .................... ....................
W 1587 hCoV-19/United Arab Emirates/0600/2020|EPI_ISL_698 .................... .................... ..................... .................... .................... ....................
W 1588 hCoV-19/United Arab Emirates/0601/2020|EPI_ISL_698 .................... .................... ..................... .................... .................... ....................
W 1589 hCoV-19/United Arab Emirates/0602/2020|EPI_ISL_698 .................... .................... ..................... .................... .................... ....................
W 1590 hCoV-19/United Arab Emirates/0603/2020|EPI_ISL_698 .................... .................... ..................... .................... .................... ....................
W 1591 hCoV-19/United Arab Emirates/0605/2020|EPI_ISL_698 .................... .................... ..................... .................... .................... ....................
W 1592 hCoV-19/United Arab Emirates/0607/2020|EPI_ISL_698 .................... .................... ..................... .................... .................... ....................
W 1593 hCoV-19/United Arab Emirates/0608/2020|EPI_ISL_698 .................... .................... ..................... .................... .................... ....................
W 1594 hCoV-19/United Arab Emirates/0609/2020|EPI_ISL_698 .................... .................... ..................... .................... .................... ....................
W 1595 hCoV-19/United Arab Emirates/0610/2020|EPI_ISL_698 .................... .................... ..................... .................... .................... ....................
W 1596 hCoV-19/United Arab Emirates/0611/2020|EPI_ISL_698 .................... .................... ..................... .................... .................... ....................
W 1597 hCoV-19/United Arab Emirates/0612/2020|EPI_ISL_698 .................... .................... ..................... .................... .................... ....................
W 1598 hCoV-19/United Arab Emirates/0613/2020|EPI_ISL_698 .................... .................... ..................... .................... .................... ....................
W 1599 hCoV-19/United Arab Emirates/0614/2020|EPI_ISL_698 .................... .................... ..................... .................... .................... ....................
W 1600 hCoV-19/United Arab Emirates/0615/2020|EPI_ISL_698 .................... .................... ..................... .................... .................... ....................
W 1601 hCoV-19/United Arab Emirates/0616/2020|EPI_ISL_698 .................... .................... ..................... .................... .................... ....................
W 1602 hCoV-19/United Arab Emirates/0617/2020|EPI_ISL_698 .................... .................... ..................... .................... .................... ....................
W 1603 hCoV-19/United Arab Emirates/0618/2020|EPI_ISL_698 .................... .................... ..................... .................... .................... ....................
W 1604 hCoV-19/United Arab Emirates/0619/2020|EPI_ISL_698 .................... .................... ..................... .................... .................... ....................
W 1605 hCoV-19/United Arab Emirates/0620/2020|EPI_ISL_698 .................... .................... ..................... .................... .................... ....................
W 1606 hCoV-19/United Arab Emirates/0621/2020|EPI_ISL_698 .................... .................... ..................... .................... .................... ....................
W 1607 hCoV-19/United Arab Emirates/0622/2020|EPI_ISL_698 .................... .................... ..................... .................... .................... ....................
W 1608 hCoV-19/United Arab Emirates/0623/2020|EPI_ISL_698 .................... .................... ..................... .................... .................... ....................
W 1609 hCoV-19/United Arab Emirates/0624/2020|EPI_ISL_698 .................... .................... ..................... .................... .................... ....................
W 1610 hCoV-19/United Arab Emirates/0625/2020|EPI_ISL_698 .................... .................... ..................... .................... .................... ....................
W 1611 hCoV-19/United Arab Emirates/0626/2020|EPI_ISL_698 .................... .................... ..................... .................... .................... ....................
W 1612 hCoV-19/United Arab Emirates/0627/2020|EPI_ISL_698 .................... .................... ..................... .................... .................... ....................
W 1613 hCoV-19/United Arab Emirates/0628/2020|EPI_ISL_698 .................... .................... ..................... .................... .................... ....................
W 1614 hCoV-19/United Arab Emirates/0629/2020|EPI_ISL_698 .................... .................... ..................... .................... .................... ....................

| | | | | | | | |
|---|---|---|---|---|---|---|---|
| W | 1615 hCoV-19/United Arab Emirates/0630/2020\|EPI_ISL_698 | ···················· | ···················· | ···················· | ···················· | ···················· | ···················· |
| W | 1616 hCoV-19/United Arab Emirates/0631/2020\|EPI_ISL_698 | ···················· | ···················· | ···················· | ···················· | ···················· | ···················· |
| W | 1617 hCoV-19/United Arab Emirates/0632/2020\|EPI_ISL_698 | ···················· | ···················· | ···················· | ···················· | ···················· | ···················· |
| W | 1618 hCoV-19/United Arab Emirates/0633/2020\|EPI_ISL_698 | ···················· | ···················· | ···················· | ···················· | ···················· | ···················· |
| W | 1619 hCoV-19/United Arab Emirates/0634/2020\|EPI_ISL_698 | ···················· | ···················· | ···················· | ···················· | ···················· | ···················· |
| W | 1620 hCoV-19/United Arab Emirates/0635/2020\|EPI_ISL_698 | ···················· | ···················· | ···················· | ···················· | ···················· | ···················· |
| W | 1621 hCoV-19/United Arab Emirates/0636/2020\|EPI_ISL_698 | ···················· | ···················· | ···················· | ···················· | ···················· | ···················· |
| W | 1622 hCoV-19/United Arab Emirates/0637/2020\|EPI_ISL_698 | ···················· | ···················· | ···················· | ···················· | ···················· | ···················· |
| W | 1623 hCoV-19/United Arab Emirates/0638/2020\|EPI_ISL_698 | ···················· | ···················· | ···················· | ···················· | ···················· | ···················· |
| W | 1624 hCoV-19/United Arab Emirates/0639/2020\|EPI_ISL_698 | ···················· | ···················· | ···················· | ···················· | ···················· | ···················· |
| W | 1625 hCoV-19/United Arab Emirates/0640/2020\|EPI_ISL_698 | ···················· | ···················· | ···················· | ···················· | ···················· | ···················· |
| W | 1626 hCoV-19/United Arab Emirates/0641/2020\|EPI_ISL_698 | ···················· | ···················· | ···················· | ···················· | ···················· | ···················· |
| W | 1627 hCoV-19/United Arab Emirates/0642/2020\|EPI_ISL_698 | ···················· | ···················· | ···················· | ···················· | ···················· | ···················· |
| W | 1629 hCoV-19/South Africa/NHLS-UCT-GS-1407/2020\|EPI_ISL | ···················· | ·A·················· | ···················· | ···················· | ···················· | ···················· |
| W | 1631 hCoV-19/South Africa/NHLS-UCT-GP-5482/2020\|EPI_ISL | ···················· | ···················· | ···················· | ···················· | ···················· | ···················· |
| W | 1633 hCoV-19/South Africa/NHLS-UCT-GP-5257/2020\|EPI_ISL | ···················· | ·A·················· | ···················· | ···················· | ···················· | ···················· |
| W | 1634 hCoV-19/South Africa/NHLS-UCT-GP-5276/2020\|EPI_ISL | ···················· | ···················· | ···················· | ···················· | ···················· | ···················· |
| W | 1635 hCoV-19/South Africa/NHLS-UCT-GP-5557/2020\|EPI_ISL | ···················· | ·A·················· | ···················· | ···················· | ···················· | ···················· |
| W | 1636 hCoV-19/South Africa/NHLS-UCT-GP-5312/2020\|EPI_ISL | ···················· | ···················· | ···················· | ···················· | ···················· | ···················· |
| W | 1637 hCoV-19/South Africa/NHLS-UCT-GP-5478/2020\|EPI_ISL | ···················· | ···················· | ···················· | ···················· | ···················· | ···················· |
| W | 1638 hCoV-19/South Africa/NHLS-UCT-GP-5671/2020\|EPI_ISL | ···················· | ·A·················· | ···················· | ···················· | ···················· | ···················· |
| W | 1639 hCoV-19/South Africa/NHLS-UCT-GP-5855/2020\|EPI_ISL | ···················· | ·A·················· | ···················· | ···················· | ···················· | ···················· |
| W | 1640 hCoV-19/South Africa/NHLS-UCT-GS-0951/2020\|EPI_ISL | ···················· | ·A·················· | ···················· | ···················· | ···················· | ···················· |
| W | 1641 hCoV-19/South Africa/NHLS-UCT-GS-0958/2020\|EPI_ISL | ···················· | ·A·················· | ···················· | ···················· | ···················· | ···················· |
| W | 1643 hCoV-19/South Africa/NHLS-UCT-GS-2211/2020\|EPI_ISL | ···················· | ·A·················· | ···················· | ···················· | ···················· | ···················· |
| W | 1644 hCoV-19/South Africa/NHLS-UCT-GP-5231/2020\|EPI_ISL | ···················· | ···················· | ···················· | ···················· | ···················· | ···················· |
| W | 1645 hCoV-19/South Africa/NHLS-UCT-GP-5411/2020\|EPI_ISL | ···················· | ···················· | ···················· | ···················· | ···················· | ···················· |
| W | 1647 hCoV-19/South Africa/NHLS-UCT-GP-5269/2020\|EPI_ISL | ···················· | ·A·················· | ···················· | ···················· | ···················· | ···················· |
| W | 1648 hCoV-19/England/ALDP-BB3AA9/2020\|EPI_ISL_701800\|20 | ···················· | ···················· | ···················· | ···················· | ···················· | ···················· |
| W | 1649 hCoV-19/England/ALDP-BB53AC/2020\|EPI_ISL_701802\|20 | ···················· | ···················· | ···················· | ···················· | ···················· | ···················· |
| W | 1650 hCoV-19/England/ALDP-BB9E01/2020\|EPI_ISL_701804\|20 | ···················· | ···················· | ···················· | ···················· | ···················· | ···················· |
| W | 1651 hCoV-19/England/ALDP-BBB0EB/2020\|EPI_ISL_701805\|20 | ···················· | ···················· | ···················· | ···················· | ···················· | ···················· |
| W | 1652 hCoV-19/England/ALDP-BBA8C6/2020\|EPI_ISL_701806\|20 | ···················· | ···················· | ···················· | ···················· | ···················· | ···················· |
| W | 1653 hCoV-19/England/ALDP-BB4F9A/2020\|EPI_ISL_701807\|20 | ···················· | ···················· | ···················· | ···················· | ···················· | ···················· |
| W | 1654 hCoV-19/England/ALDP-BBA40E/2020\|EPI_ISL_701808\|20 | ···················· | ···················· | ···················· | ···················· | ···················· | ···················· |
| W | 1655 hCoV-19/England/ALDP-BB3DBF/2020\|EPI_ISL_701809\|20 | ···················· | ···················· | ···················· | ···················· | ···················· | ···················· |
| W | 1656 hCoV-19/England/ALDP-BB51EE/2020\|EPI_ISL_701810\|20 | ···················· | ···················· | ···················· | ···················· | ···················· | ···················· |
| W | 1657 hCoV-19/England/ALDP-BB3D46/2020\|EPI_ISL_701811\|20 | ···················· | ···················· | ···················· | ···················· | ···················· | ···················· |
| W | 1658 hCoV-19/England/ALDP-BBB29A/2020\|EPI_ISL_701813\|20 | ···················· | ···················· | ···················· | ···················· | ···················· | ···················· |
| W | 1659 hCoV-19/England/ALDP-BBA8A8/2020\|EPI_ISL_701814\|20 | ···················· | ···················· | ···················· | ···················· | ···················· | ···················· |
| W | 1660 hCoV-19/England/ALDP-BB9E89/2020\|EPI_ISL_701815\|20 | ···················· | ···················· | ···················· | ···················· | ···················· | ···················· |
| W | 1661 hCoV-19/England/ALDP-BBA116/2020\|EPI_ISL_701817\|20 | ···················· | ···················· | ···················· | ···················· | ···················· | ···················· |
| W | 1662 hCoV-19/England/ALDP-BB9F0E/2020\|EPI_ISL_701818\|20 | ···················· | ···················· | ···················· | ···················· | ···················· | ···················· |
| W | 1663 hCoV-19/England/ALDP-BB38BE/2020\|EPI_ISL_701819\|20 | ···················· | ···················· | ···················· | ···················· | ···················· | ···················· |
| W | 1664 hCoV-19/England/ALDP-BBB072/2020\|EPI_ISL_701820\|20 | ···················· | ···················· | ···················· | ···················· | ···················· | ···················· |
| W | 1665 hCoV-19/England/ALDP-BBA78D/2020\|EPI_ISL_701821\|20 | ···················· | ···················· | ···················· | ···················· | ···················· | ···················· |
| W | 1666 hCoV-19/England/ALDP-BB5087/2020\|EPI_ISL_701822\|20 | ···················· | ···················· | ···················· | ···················· | ···················· | ···················· |
| W | 1667 hCoV-19/England/ALDP-BBB25E/2020\|EPI_ISL_701823\|20 | ···················· | ···················· | ···················· | ···················· | ···················· | ···················· |
| W | 1668 hCoV-19/England/ALDP-BBA741/2020\|EPI_ISL_701824\|20 | ···················· | ···················· | ···················· | ···················· | ···················· | ···················· |
| W | 1669 hCoV-19/England/ALDP-BB51B1/2020\|EPI_ISL_701825\|20 | ···················· | ···················· | ···················· | ···················· | ···················· | ···················· |
| W | 1670 hCoV-19/England/ALDP-BB9EB6/2020\|EPI_ISL_701826\|20 | ···················· | ···················· | ···················· | ···················· | ···················· | ···················· |
| W | 1671 hCoV-19/England/ALDP-BB5245/2020\|EPI_ISL_701827\|20 | ···················· | ···················· | ···················· | ···················· | ···················· | ···················· |
| W | 1672 hCoV-19/England/ALDP-BB3C58/2020\|EPI_ISL_701828\|20 | ···················· | ···················· | ···················· | ···················· | ···················· | ···················· |
| W | 1673 hCoV-19/England/ALDP-BB50C3/2020\|EPI_ISL_701829\|20 | ···················· | ···················· | ···················· | ···················· | ···················· | ···················· |
| W | 1674 hCoV-19/England/ALDP-BBB528/2020\|EPI_ISL_701831\|20 | ···················· | ···················· | ···················· | ···················· | ···················· | ···················· |
| W | 1675 hCoV-19/England/ALDP-BB501E/2020\|EPI_ISL_701832\|20 | ···················· | ···················· | ···················· | ···················· | ···················· | ···················· |
| W | 1676 hCoV-19/England/ALDP-BB3D73/2020\|EPI_ISL_701833\|20 | ···················· | ···················· | ···················· | ···················· | ···················· | ···················· |
| W | 1677 hCoV-19/England/ALDP-BBB106/2020\|EPI_ISL_701834\|20 | ···················· | ···················· | ···················· | ···················· | ···················· | ···················· |
| W | 1678 hCoV-19/England/ALDP-BBA85D/2020\|EPI_ISL_701859\|20 | ···················· | ···················· | ···················· | ···················· | ···················· | ···················· |
| W | 1679 hCoV-19/England/ALDP-BBA41D/2020\|EPI_ISL_701860\|20 | ············T······· | ···················· | ···················· | ···················· | ···················· | ···················· |
| W | 1680 hCoV-19/England/ALDP-BB561C/2020\|EPI_ISL_701861\|20 | ···················· | ···················· | ···················· | ···················· | ···················· | ···················· |
| W | 1681 hCoV-19/England/ALDP-BBB4FE/2020\|EPI_ISL_701862\|20 | ···················· | ···················· | ···················· | ···················· | ···················· | ···················· |
| W | 1682 hCoV-19/England/ALDP-BB3933/2020\|EPI_ISL_701865\|20 | ···················· | ···················· | ···················· | ···················· | ···················· | ···················· |
| W | 1683 hCoV-19/England/ALDP-BBAC24/2020\|EPI_ISL_701866\|20 | ···················· | ···················· | ···················· | ···················· | ···················· | ···················· |
| W | 1684 hCoV-19/England/ALDP-BBB0FA/2020\|EPI_ISL_701867\|20 | ···················· | ···················· | ···················· | ···················· | ···················· | ···················· |
| W | 1685 hCoV-19/England/ALDP-BBB009/2020\|EPI_ISL_701869\|20 | ···················· | ···················· | ···················· | ···················· | ···················· | ···················· |
| W | 1686 hCoV-19/England/ALDP-BB3C1C/2020\|EPI_ISL_701871\|20 | ···················· | ···················· | ···················· | ···················· | ···················· | ···················· |
| W | 1687 hCoV-19/England/ALDP-BB5430/2020\|EPI_ISL_701872\|20 | ···················· | ···················· | ···················· | ···················· | ···················· | ···················· |
| W | 1688 hCoV-19/England/ALDP-BBA3D4/2020\|EPI_ISL_701873\|20 | ···················· | ···················· | ···················· | ···················· | ···················· | ···················· |
| W | 1689 hCoV-19/England/QEUH-BC39E5/2020\|EPI_ISL_704000\|20 | ···················· | ···················· | ···················· | ···················· | ···················· | ···················· |
| W | 1690 hCoV-19/England/QEUH-BC3B49/2020\|EPI_ISL_704002\|20 | ···················· | ···················· | ···················· | ···················· | ···················· | ···················· |
| W | 1691 hCoV-19/England/QEUH-BC4B66/2020\|EPI_ISL_704004\|20 | ···················· | ···················· | ···················· | ···················· | ···················· | ···················· |
| W | 1692 hCoV-19/England/QEUH-BC3BEF/2020\|EPI_ISL_704005\|20 | ···················· | ·A·················· | ···················· | ···················· | ···················· | ···A················ |
| W | 1693 hCoV-19/England/QEUH-BC3790/2020\|EPI_ISL_704007\|20 | ···················· | ···················· | ···················· | ···················· | ···················· | ···················· |
| W | 1694 hCoV-19/England/QEUH-BC4AE1/2020\|EPI_ISL_704008\|20 | ···················· | ···················· | ···················· | ···················· | ···················· | ···················· |
| W | 1695 hCoV-19/England/QEUH-BC4B75/2020\|EPI_ISL_704018\|20 | ···················· | ···················· | ···················· | ···················· | ···················· | ···················· |
| W | 1696 hCoV-19/England/QEUH-BC4A69/2020\|EPI_ISL_704023\|20 | ···················· | ···················· | ···················· | ···················· | ···················· | ···················· |
| W | 1697 hCoV-19/England/QEUH-BC3745/2020\|EPI_ISL_704024\|20 | ···················· | ···················· | ···················· | ···················· | ···················· | ···················· |
| W | 1698 hCoV-19/England/QEUH-BCC779/2020\|EPI_ISL_704026\|20 | ···················· | ···················· | ···················· | ···················· | ···················· | ···················· |
| W | 1699 hCoV-19/England/QEUH-BCC269/2020\|EPI_ISL_704027\|20 | ···················· | ···················· | ···················· | ···················· | ···················· | ···················· |

| | | | | | | | |
|---|---|---|---|---|---|---|---|
| W | 1700 hCoV-19/England/QEUH-BCC111/2020\|EPI_ISL_704029\|20 | ···················· | ···················· | ····················· | ···················· | ···················· | ···················· |
| W | 1701 hCoV-19/England/QEUH-BCC74C/2020\|EPI_ISL_704031\|20 | ···················· | ···················· | ····················· | ···················· | ···················· | ···················· |
| W | 1702 hCoV-19/England/QEUH-BC3AB5/2020\|EPI_ISL_704032\|20 | ···················· | ···················· | ····················· | ···················· | ···················· | ···················· |
| W | 1703 hCoV-19/England/QEUH-BCB8D1/2020\|EPI_ISL_704034\|20 | ···················· | ···················· | ····················· | ···················· | ···················· | ···················· |
| W | 1704 hCoV-19/England/QEUH-BCBE39/2020\|EPI_ISL_704035\|20 | ···················· | ···················· | ····················· | ···················· | ···················· | ···················· |
| W | 1705 hCoV-19/England/QEUH-BCB859/2020\|EPI_ISL_704037\|20 | ···················· | ···················· | ····················· | ···················· | ···················· | ···················· |
| W | 1706 hCoV-19/England/QEUH-BCBD2D/2020\|EPI_ISL_704040\|20 | ···················· | ···················· | ····················· | ···················· | ·····T·············· | ···················· |
| W | 1707 hCoV-19/England/QEUH-BCC603/2020\|EPI_ISL_704042\|20 | ···················· | ···················· | ····················· | ···················· | ···················· | ···················· |
| W | 1708 hCoV-19/England/QEUH-BCBD87/2020\|EPI_ISL_704045\|20 | ···················· | ···················· | ····················· | ···················· | ···················· | ···················· |
| W | 1709 hCoV-19/England/QEUH-BCC09C/2020\|EPI_ISL_704047\|20 | ···················· | ···················· | ····················· | ···················· | ···················· | ···················· |
| W | 1710 hCoV-19/England/QEUH-BC360C/2020\|EPI_ISL_704048\|20 | ···················· | ···················· | ····················· | ···················· | ···················· | ···················· |
| W | 1711 hCoV-19/England/QEUH-BC3A97/2020\|EPI_ISL_704051\|20 | ···················· | ···················· | ····················· | ···················· | ···················· | ···················· |
| W | 1712 hCoV-19/England/QEUH-BCB455/2020\|EPI_ISL_704053\|20 | ···················· | ·A·················· | ····················· | ···················· | ···················· | ···················· |
| W | 1713 hCoV-19/England/QEUH-BCC5BB/2020\|EPI_ISL_704055\|20 | ···················· | ···················· | ····················· | ···················· | ···················· | ···················· |
| W | 1714 hCoV-19/England/QEUH-BCC6D6/2020\|EPI_ISL_704056\|20 | ···················· | ···················· | ····················· | ···················· | ···················· | ···················· |
| W | 1715 hCoV-19/England/QEUH-BCBCE4/2020\|EPI_ISL_704058\|20 | ···················· | ···················· | ····················· | ···················· | ···················· | ···················· |
| W | 1716 hCoV-19/England/QEUH-BC4AB4/2020\|EPI_ISL_704059\|20 | ···················· | ···················· | ····················· | ···················· | ···················· | ···················· |
| W | 1717 hCoV-19/England/QEUH-BC37AF/2020\|EPI_ISL_704061\|20 | ···················· | ···················· | ····················· | ···················· | ···················· | ···················· |
| W | 1718 hCoV-19/England/QEUH-BC37DC/2020\|EPI_ISL_704063\|20 | ···················· | ···················· | ····················· | ···················· | ···················· | ···················· |
| W | 1719 hCoV-19/England/QEUH-BCB69B/2020\|EPI_ISL_704064\|20 | ···················· | ···················· | ····················· | ···················· | ···················· | ···················· |
| W | 1720 hCoV-19/England/QEUH-BC4A4B/2020\|EPI_ISL_704066\|20 | ···················· | ···················· | ····················· | ···················· | ···················· | ···················· |
| W | 1721 hCoV-19/England/QEUH-BCBA35/2020\|EPI_ISL_704067\|20 | ···················· | ···················· | ····················· | ···················· | ···················· | ···················· |
| W | 1722 hCoV-19/England/QEUH-BCB90B/2020\|EPI_ISL_704071\|20 | ···················· | ···················· | ····················· | ···················· | ···················· | ···················· |
| W | 1723 hCoV-19/England/QEUH-BC3AF1/2020\|EPI_ISL_704072\|20 | ···················· | ···················· | ····················· | ···················· | ···················· | ···················· |
| W | 1724 hCoV-19/England/QEUH-BCC0BA/2020\|EPI_ISL_704077\|20 | ···················· | ···················· | ····················· | ···················· | ···················· | ···················· |
| W | 1725 hCoV-19/England/QEUH-BC4A3C/2020\|EPI_ISL_704078\|20 | ···················· | ···················· | ····················· | ···················· | ···················· | ···················· |
| W | 1726 hCoV-19/England/QEUH-BC4A0F/2020\|EPI_ISL_704080\|20 | ···················· | ···················· | ····················· | ···················· | ···················· | ···················· |
| W | 1727 hCoV-19/England/QEUH-BCC0AB/2020\|EPI_ISL_704082\|20 | ···················· | ···················· | ····················· | ···················· | ·····T·············· | ···················· |
| W | 1728 hCoV-19/England/QEUH-BC4AF0/2020\|EPI_ISL_704083\|20 | ···················· | ···················· | ····················· | ···················· | ···················· | ···················· |
| W | 1729 hCoV-19/England/QEUH-BC3718/2020\|EPI_ISL_704085\|20 | ···················· | ···················· | ····················· | ···················· | ···················· | ···················· |
| W | 1730 hCoV-19/England/QEUH-BCBD0F/2020\|EPI_ISL_704088\|20 | ···················· | ···················· | ····················· | ···················· | ···················· | ···················· |
| W | 1731 hCoV-19/England/QEUH-BCBF81/2020\|EPI_ISL_704090\|20 | ···················· | ···················· | ····················· | ···················· | ···················· | ···················· |
| W | 1732 hCoV-19/England/QEUH-BC3B94/2020\|EPI_ISL_704091\|20 | ···················· | ···················· | ····················· | ···················· | ···················· | ···················· |
| W | 1733 hCoV-19/England/QEUH-BC4A87/2020\|EPI_ISL_704093\|20 | ···················· | ···················· | ····················· | ···················· | ···················· | ···················· |
| W | 1734 hCoV-19/England/QEUH-BC4B93/2020\|EPI_ISL_704094\|20 | ···················· | ···················· | ····················· | ···················· | ···················· | ···················· |
| W | 1735 hCoV-19/England/QEUH-BC3BD0/2020\|EPI_ISL_704096\|20 | ···················· | ···················· | ····················· | ···················· | ···················· | ···················· |
| W | 1736 hCoV-19/England/QEUH-BC36A2/2020\|EPI_ISL_704098\|20 | ···················· | ···················· | ····················· | ···················· | ·····T·············· | ···················· |
| W | 1737 hCoV-19/England/QEUH-BCB895/2020\|EPI_ISL_704099\|20 | ···················· | ···················· | ····················· | ···················· | ···················· | ···················· |
| W | 1738 hCoV-19/Wales/PHWC-4913E9/2020\|EPI_ISL_706650\|2020 | ···················· | ···················· | ····················· | ···················· | ···················· | ···················· |
| W | 1742 hCoV-19/Wales/PHWC-491440/2020\|EPI_ISL_706654\|2020 | ···················· | ···················· | ····················· | ···················· | ···················· | ···················· |
| W | 1743 hCoV-19/Wales/PHWC-49145F/2020\|EPI_ISL_706655\|2020 | ···················· | ···················· | ····················· | ···················· | ···················· | ···················· |
| W | 1745 hCoV-19/Wales/PHWC-49148C/2020\|EPI_ISL_706657\|2020 | ···················· | ···················· | ····················· | ···················· | ···················· | ···················· |
| W | 1746 hCoV-19/Wales/PHWC-49149B/2020\|EPI_ISL_706658\|2020 | ···················· | ···················· | ····················· | ···················· | ···················· | NN·················· |
| W | 1748 hCoV-19/Wales/PHWC-4914B9/2020\|EPI_ISL_706660\|2020 | ···················· | ···················· | ····················· | ···················· | ···················· | ···················· |
| W | 1749 hCoV-19/Wales/PHWC-4914C8/2020\|EPI_ISL_706661\|2020 | ···················· | ···················· | ····················· | ···················· | ···················· | ···················· |
| W | 1750 hCoV-19/Wales/PHWC-4914F5/2020\|EPI_ISL_706662\|2020 | ···················· | ······A············· | ····················· | ···················· | ···················· | ···················· |
| W | 1758 hCoV-19/Wales/PHWC-4916A4/2020\|EPI_ISL_706678\|2020 | ···················· | ···················· | ····················· | ···················· | ···················· | ···················· |
| W | 1760 hCoV-19/Wales/PHWC-4916C2/2020\|EPI_ISL_706680\|2020 | ···················· | ···················· | ····················· | ···················· | ···················· | ···················· |
| W | 1761 hCoV-19/Wales/PHWC-4916E0/2020\|EPI_ISL_706681\|2020 | ···················· | ···················· | ····················· | ···················· | ···················· | ···················· |
| W | 1763 hCoV-19/Wales/PHWC-49170B/2020\|EPI_ISL_706683\|2020 | ···················· | ···················· | ····················· | ···················· | ···················· | ···················· |
| W | 1764 hCoV-19/Wales/PHWC-49171A/2020\|EPI_ISL_706684\|2020 | ···················· | ···················· | ····················· | ···················· | ···················· | ···················· |
| W | 1766 hCoV-19/Wales/PHWC-491738/2020\|EPI_ISL_706686\|2020 | ···················· | ···················· | ····················· | ···················· | ···················· | ···················· |
| W | 1769 hCoV-19/Wales/PHWC-491765/2020\|EPI_ISL_706689\|2020 | ···················· | ···················· | ····················· | ···················· | ···················· | ···················· |
| W | 1770 hCoV-19/Wales/PHWC-491774/2020\|EPI_ISL_706690\|2020 | ···················· | ···················· | ····················· | ···················· | ···················· | ···················· |
| W | 1771 hCoV-19/Wales/PHWC-491783/2020\|EPI_ISL_706691\|2020 | ···················· | ···················· | ····················· | ···················· | ···················· | ···················· |
| W | 1772 hCoV-19/Wales/PHWC-491792/2020\|EPI_ISL_706692\|2020 | ···················· | ···················· | ····················· | ···················· | ···················· | ···················· |
| W | 1774 hCoV-19/Wales/PHWC-4917CF/2020\|EPI_ISL_706694\|2020 | ···················· | ···················· | ····················· | ···················· | ···················· | ···················· |
| W | 1775 hCoV-19/Wales/PHWC-4917ED/2020\|EPI_ISL_706695\|2020 | ···················· | ···················· | ····················· | ···················· | ···················· | ···················· |
| W | 1776 hCoV-19/Wales/PHWC-4917FC/2020\|EPI_ISL_706696\|2020 | ···················· | ···················· | ····················· | ···················· | ···················· | ···················· |
| W | 1778 hCoV-19/Wales/PHWC-491826/2020\|EPI_ISL_706698\|2020 | ···················· | ···················· | ····················· | ···················· | ···················· | ···················· |
| W | 1779 hCoV-19/Wales/PHWC-491835/2020\|EPI_ISL_706699\|2020 | ···················· | ···················· | ····················· | ···················· | ···················· | ···················· |
| W | 1780 hCoV-19/Germany/SL-SU-10450660/2020\|EPI_ISL_707981 | ···················· | ···················· | ····················· | ···················· | ···················· | ···················· |
| W | 1781 hCoV-19/Germany/SL-SU-10450668/2020\|EPI_ISL_707982 | ···················· | ···················· | ····················· | ···················· | ···················· | ···················· |
| W | 1782 hCoV-19/Germany/SL-SU-10450986/2020\|EPI_ISL_707986 | ···················· | ···················· | ····················· | ···················· | ···················· | ···················· |
| W | 1783 hCoV-19/Germany/SL-SU-10451157/2020\|EPI_ISL_707987 | ···················· | ···················· | ····················· | ···················· | ···················· | ···················· |
| W | 1784 hCoV-19/Germany/SL-SU-10451160/2020\|EPI_ISL_707988 | ···················· | ···················· | ····················· | ···················· | ···················· | ···················· |
| W | 1785 hCoV-19/Germany/SL-SU-10451375/2020\|EPI_ISL_707991 | ···················· | ···················· | ····················· | ···················· | ···················· | ···················· |
| W | 1786 hCoV-19/Germany/SL-SU-10451472/2020\|EPI_ISL_707992 | ···················· | ···················· | ····················· | ···················· | ···················· | ···················· |
| W | 1787 hCoV-19/Germany/SL-SU-10451495/2020\|EPI_ISL_707993 | ···················· | ···················· | ····················· | ···················· | ···················· | ···················· |
| W | 1789 hCoV-19/Germany/SL-SU-10451904/2020\|EPI_ISL_707995 | ···················· | ···················· | ····················· | ···················· | ···················· | ···················· |
| W | 1790 hCoV-19/Germany/SL-SU-10452029/2020\|EPI_ISL_707996 | ···················· | ···················· | ····················· | ···················· | ·····T·············· | ···················· |
| W | 1791 hCoV-19/Germany/SL-SU-10452037/2020\|EPI_ISL_707998 | ···················· | ···················· | ····················· | ···················· | ···················· | ···················· |
| W | 1792 hCoV-19/Germany/SL-SU-10452356/2020\|EPI_ISL_707999 | ···················· | ···················· | ····················· | ···················· | ···················· | ···················· |
| W | 1793 hCoV-19/USA/AZ-TG654712/2020\|EPI_ISL_696018\|2020-0 | ···················· | ···················· | ····················· | ···················· | ···················· | ···················· |
| W | 1794 hCoV-19/USA/AZ-TG654719/2020\|EPI_ISL_696019\|2020-0 | ···················· | ···················· | ····················· | ···················· | ···················· | ···················· |
| W | 1795 hCoV-19/USA/AZ-TG654730/2020\|EPI_ISL_696020\|2020-0 | ···················· | ···················· | ····················· | ···················· | ···················· | ···················· |
| W | 1796 hCoV-19/USA/AZ-TG654739/2020\|EPI_ISL_696021\|2020-0 | ···················· | ···················· | ····················· | ···················· | ···················· | ···················· |
| W | 1797 hCoV-19/USA/AZ-TG654743/2020\|EPI_ISL_696022\|2020-0 | ···················· | ···················· | ····················· | ···················· | ···················· | ···················· |
| W | 1798 hCoV-19/USA/AZ-TG654747/2020\|EPI_ISL_696023\|2020-0 | ···················· | ···················· | ····················· | ···················· | ···················· | ···················· |
| W | 1799 hCoV-19/USA/AZ-TG664219/2020\|EPI_ISL_696028\|2020-0 | ···················· | ···················· | ····················· | ···················· | ···················· | ···················· |

| | | | | | | | | |
|---|---|---|---|---|---|---|---|---|
| W | 1800 hCoV-19/USA/AZ-TG664220/2020\|EPI_ISL_696029\|2020-0 | ···················· | ···················· | ···················· | ···················· | ···················· | ···················· | |
| W | 1801 hCoV-19/USA/AZ-TG664226/2020\|EPI_ISL_696034\|2020-0 | ···················· | ···················· | ···················· | ···················· | ···················· | ···················· | |
| W | 1802 hCoV-19/USA/AZ-TG664228/2020\|EPI_ISL_696036\|2020-0 | ···················· | ···················· | ···················· | ···················· | ···················· | ···················· | |
| W | 1803 hCoV-19/USA/AZ-TG664239/2020\|EPI_ISL_696043\|2020-0 | ···················· | ···················· | ···················· | ···················· | ···················· | ···················· | |
| W | 1804 hCoV-19/USA/AZ-TG664243/2020\|EPI_ISL_696047\|2020-0 | ···················· | ···················· | ···················· | ···················· | ···················· | ···················· | |
| W | 1805 hCoV-19/USA/AZ-TG664244/2020\|EPI_ISL_696048\|2020-0 | ···················· | ···················· | ···················· | ···················· | ···················· | ···················· | |
| W | 1806 hCoV-19/USA/AZ-TG664246/2020\|EPI_ISL_696049\|2020-0 | ···················· | ···················· | ···················· | ···················· | ···················· | ···················· | |
| W | 1807 hCoV-19/USA/AZ-TG664248/2020\|EPI_ISL_696051\|2020-0 | ···················· | ···················· | ···················· | ···················· | ···················· | ···················· | |
| W | 1808 hCoV-19/USA/AZ-TG664258/2020\|EPI_ISL_696060\|2020-0 | ···················· | ···················· | ···················· | ···················· | ···················· | ···················· | |
| W | 1809 hCoV-19/USA/AZ-TG664264/2020\|EPI_ISL_696065\|2020-0 | ···················· | ···················· | ···················· | ···················· | ···················· | ···················· | |
| W | 1810 hCoV-19/USA/AZ-TG664275/2020\|EPI_ISL_696076\|2020-0 | ···················· | ···················· | ···················· | ···················· | ···················· | ···················· | |
| W | 1811 hCoV-19/USA/AZ-TG664276/2020\|EPI_ISL_696077\|2020-0 | ···················· | ···················· | ···················· | ···················· | ···················· | ···················· | |
| W | 1812 hCoV-19/USA/AZ-TG664277/2020\|EPI_ISL_696078\|2020-0 | ···················· | ···················· | ···················· | ···················· | ···················· | ···················· | |
| W | 1813 hCoV-19/USA/AZ-TG664281/2020\|EPI_ISL_696082\|2020-0 | ···················· | ···················· | ···················· | ···················· | ···················· | ···················· | |
| W | 1814 hCoV-19/United Arab Emirates/0515/2020\|EPI_ISL_698 | ···················· | ···················· | ···················· | ···················· | ···················· | ···················· | |
| W | 1815 hCoV-19/United Arab Emirates/0516/2020\|EPI_ISL_698 | ···················· | ···················· | ···················· | ···················· | ···················· | ···················· | |
| W | 1816 hCoV-19/United Arab Emirates/0517/2020\|EPI_ISL_698 | ···················· | ···················· | ···················· | ···················· | ···················· | ···················· | |
| W | 1817 hCoV-19/United Arab Emirates/0518/2020\|EPI_ISL_698 | ···················· | ···················· | ···················· | ···················· | ···················· | ···················· | |
| W | 1818 hCoV-19/United Arab Emirates/0519/2020\|EPI_ISL_698 | ···················· | ···················· | ···················· | ···················· | ···················· | ···················· | |
| W | 1819 hCoV-19/United Arab Emirates/0520/2020\|EPI_ISL_698 | ···················· | ···················· | ···················· | ···················· | ···················· | ···················· | |
| W | 1820 hCoV-19/United Arab Emirates/0521/2020\|EPI_ISL_698 | ···················· | ···················· | ···················· | ···················· | ···················· | ···················· | |
| W | 1821 hCoV-19/United Arab Emirates/0522/2020\|EPI_ISL_698 | ···················· | ···················· | ···················· | ···················· | ···················· | ···················· | |
| W | 1822 hCoV-19/United Arab Emirates/0523/2020\|EPI_ISL_698 | ···················· | ···················· | ···················· | ···················· | ···················· | ···················· | |
| W | 1823 hCoV-19/United Arab Emirates/0524/2020\|EPI_ISL_698 | ···················· | ···················· | ···················· | ···················· | ···················· | ···················· | |
| W | 1824 hCoV-19/United Arab Emirates/0525/2020\|EPI_ISL_698 | ···················· | ···················· | ···················· | ···················· | ···················· | ···················· | |
| W | 1825 hCoV-19/United Arab Emirates/0526/2020\|EPI_ISL_698 | ···················· | ···················· | ···················· | ···················· | ···················· | ···················· | |
| W | 1826 hCoV-19/United Arab Emirates/0528/2020\|EPI_ISL_698 | ···················· | ···················· | ···················· | ···················· | ···················· | ···················· | |
| W | 1827 hCoV-19/United Arab Emirates/0529/2020\|EPI_ISL_698 | ···················· | ···················· | ···················· | ···················· | ···················· | ···················· | |
| W | 1828 hCoV-19/United Arab Emirates/0530/2020\|EPI_ISL_698 | ···················· | ···················· | ···················· | ···················· | ···················· | ···················· | |
| W | 1829 hCoV-19/United Arab Emirates/0531/2020\|EPI_ISL_698 | ···················· | ···················· | ···················· | ···················· | ···················· | ···················· | |
| W | 1830 hCoV-19/United Arab Emirates/0532/2020\|EPI_ISL_698 | ···················· | ···················· | ···················· | ···················· | ···················· | ···················· | |
| W | 1831 hCoV-19/United Arab Emirates/0533/2020\|EPI_ISL_698 | ···················· | ···················· | ···················· | ···················· | ···················· | ···················· | |
| W | 1832 hCoV-19/United Arab Emirates/0534/2020\|EPI_ISL_698 | ···················· | ···················· | ···················· | ···················· | ···················· | ···················· | |
| W | 1833 hCoV-19/United Arab Emirates/0535/2020\|EPI_ISL_698 | ···················· | ···················· | ···················· | ···················· | ···················· | ···················· | |
| W | 1834 hCoV-19/United Arab Emirates/0537/2020\|EPI_ISL_698 | ···················· | ···················· | ···················· | ···················· | ···················· | ···················· | |
| W | 1835 hCoV-19/United Arab Emirates/0538/2020\|EPI_ISL_698 | ···················· | ···················· | ···················· | ···················· | ···················· | ···················· | |
| W | 1836 hCoV-19/United Arab Emirates/0539/2020\|EPI_ISL_698 | ···················· | ···················· | ···················· | ···················· | ···················· | ···················· | |
| W | 1837 hCoV-19/United Arab Emirates/0540/2020\|EPI_ISL_698 | ···················· | ···················· | ···················· | ···················· | ···················· | ···················· | |
| W | 1838 hCoV-19/United Arab Emirates/0541/2020\|EPI_ISL_698 | ···················· | ···················· | ···················· | ···················· | ···················· | ···················· | |
| W | 1839 hCoV-19/United Arab Emirates/0542/2020\|EPI_ISL_698 | ···················· | ···················· | ···················· | ···················· | ···················· | ···················· | |
| W | 1840 hCoV-19/United Arab Emirates/0543/2020\|EPI_ISL_698 | ···················· | ···················· | ···················· | ···················· | ···················· | ···················· | |
| W | 1841 hCoV-19/United Arab Emirates/0544/2020\|EPI_ISL_698 | ···················· | ···················· | ···················· | ···················· | ···················· | ···················· | |
| W | 1842 hCoV-19/United Arab Emirates/0545/2020\|EPI_ISL_698 | ···················· | ···················· | ·········A·········· | ···················· | ···················· | ···················· | |
| W | 1843 hCoV-19/United Arab Emirates/0546/2020\|EPI_ISL_698 | ···················· | ······A············· | ···················· | ···················· | ···················· | ···················· | |
| W | 1844 hCoV-19/United Arab Emirates/0547/2020\|EPI_ISL_698 | ···················· | ···················· | ···················· | ···················· | ···················· | ···················· | |
| W | 1845 hCoV-19/United Arab Emirates/0548/2020\|EPI_ISL_698 | ···················· | ···················· | ···················· | ···················· | ···················· | ···················· | |
| W | 1846 hCoV-19/United Arab Emirates/0549/2020\|EPI_ISL_698 | ···················· | ···················· | ···················· | ···················· | ···················· | ···················· | |
| W | 1849 hCoV-19/South Africa/NHLS-UCT-GS-1455/2020\|EPI_ISL | ···················· | ·A·················· | ···················· | ···················· | ···················· | ···················· | |
| W | 1850 hCoV-19/South Africa/NHLS-UCT-GS-1695/2020\|EPI_ISL | ···················· | ·A·················· | ···················· | ···················· | ···················· | ···················· | |
| W | 1851 hCoV-19/South Africa/NHLS-UCT-GP-5326/2020\|EPI_ISL | ···················· | ···················· | ···················· | ···················· | ···················· | ···················· | |
| W | 1852 hCoV-19/South Africa/NHLS-UCT-GS-1582/2020\|EPI_ISL | ···················· | ·A·················· | ···················· | ···················· | ···················· | ···················· | |
| W | 1853 hCoV-19/South Africa/NHLS-UCT-GS-2131/2020\|EPI_ISL | ···················· | ···················· | ···················· | ···················· | ···················· | ···················· | |
| W | 1854 hCoV-19/South Africa/NHLS-UCT-GP-5020/2020\|EPI_ISL | ···················· | ···················· | ···················· | ···················· | ···················· | ···················· | |
| W | 1855 hCoV-19/South Africa/NHLS-UCT-GS-1971/2020\|EPI_ISL | ···················· | ···················· | ···················· | ···················· | ·NN················· | ···················· | |
| W | 1857 hCoV-19/South Africa/NHLS-UCT-GP-5593/2020\|EPI_ISL | ···················· | ···················· | ···················· | ···················· | ···················· | ···················· | |
| W | 1859 hCoV-19/South Africa/NHLS-UCT-GS-1870/2020\|EPI_ISL | ···················· | ·A·················· | ···················· | ···················· | ···················· | ···················· | |
| W | 1860 hCoV-19/South Africa/NHLS-UCT-GS-1027/2020\|EPI_ISL | ···················· | ·A·················· | ···················· | ···················· | ···················· | ···················· | |
| W | 1861 hCoV-19/South Africa/NHLS-UCT-GS-2215/2020\|EPI_ISL | ···················· | ···················· | ···················· | ···················· | ···················· | ···················· | |
| W | 1863 hCoV-19/South Africa/NHLS-UCT-GS-0641/2020\|EPI_ISL | ···················· | ·A·················· | ···················· | ···················· | ···················· | ···················· | |
| W | 1864 hCoV-19/South Africa/NHLS-UCT-GS-1440/2020\|EPI_ISL | ···················· | ···················· | ···················· | ···················· | ···················· | ···················· | |
| W | 1865 hCoV-19/South Africa/NHLS-UCT-GS-0657/2020\|EPI_ISL | ···················· | ·A·················· | ···················· | ···················· | ···················· | ···················· | |
| W | 1870 hCoV-19/South Africa/NHLS-UCT-GS-0828/2020\|EPI_ISL | ···················· | ···················· | ···················· | ···················· | ···················· | ···················· | |
| W | 1871 hCoV-19/South Africa/NHLS-UCT-GP-5244/2020\|EPI_ISL | ···················· | ···················· | ···················· | ···················· | ···················· | ···················· | |
| W | 1872 hCoV-19/South Africa/NHLS-UCT-GS-3230/2020\|EPI_ISL | ···················· | ·A·················· | ···················· | ···················· | ···················· | ···················· | |
| W | 1873 hCoV-19/South Africa/NHLS-UCT-GS-2208/2020\|EPI_ISL | ···················· | ···················· | ···················· | ···················· | ···················· | ···················· | |
| W | 1874 hCoV-19/South Africa/NHLS-UCT-GP-5792/2020\|EPI_ISL | ···················· | ···················· | ···················· | ···················· | ···················· | ···················· | |
| W | 1875 hCoV-19/South Africa/NHLS-UCT-GS-1353/2020\|EPI_ISL | ···················· | ·A·················· | ···················· | ···················· | ···················· | ···················· | |
| W | 1876 hCoV-19/South Africa/NHLS-UCT-GP-5895/2020\|EPI_ISL | ···················· | ·A·················· | ···················· | ···················· | ···················· | ···················· | |
| W | 1879 hCoV-19/South Africa/NHLS-UCT-GP-5023/2020\|EPI_ISL | ···················· | ·A·················· | ···················· | ···················· | ···················· | ···················· | |
| W | 1882 hCoV-19/South Africa/NHLS-UCT-GS-1485/2020\|EPI_ISL | ···················· | ·A·················· | ···················· | ···················· | ···················· | ···················· | |
| W | 1883 hCoV-19/South Africa/NHLS-UCT-GS-0636/2020\|EPI_ISL | ···················· | ·A·················· | ···················· | ···················· | ···················· | ···················· | |
| W | 1884 hCoV-19/South Africa/NHLS-UCT-GS-0663/2020\|EPI_ISL | ···················· | ·A·················· | ···················· | ···················· | ···················· | ···················· | |
| W | 1885 hCoV-19/South Africa/NHLS-UCT-GS-1821/2020\|EPI_ISL | ···················· | ·A·················· | ···················· | ···················· | ···················· | ···················· | |
| W | 1886 hCoV-19/South Africa/NHLS-UCT-GS-2058/2020\|EPI_ISL | ···················· | ···················· | ···················· | ···················· | ···················· | ···················· | |
| W | 1887 hCoV-19/South Africa/NHLS-UCT-GS-0681/2020\|EPI_ISL | ···················· | ···················· | ···················· | ···················· | ···················· | ···················· | |
| W | 1888 hCoV-19/South Africa/NHLS-UCT-GP-5689/2020\|EPI_ISL | ···················· | ···················· | ···················· | ···················· | ···················· | ···················· | |
| W | 1889 hCoV-19/South Africa/NHLS-UCT-GP-5889/2020\|EPI_ISL | ···················· | ···················· | ···················· | ···················· | ···················· | ···················· | |
| W | 1891 hCoV-19/South Africa/NHLS-UCT-GS-0620/2020\|EPI_ISL | ·················T·· | ···················· | ···················· | ···················· | ···················· | ···················· | 1 |
| W | 1892 hCoV-19/South Africa/NHLS-UCT-GS-1486/2020\|EPI_ISL | ···················· | ·A·················· | ···················· | ···················· | ···················· | ···················· | |
| W | 1893 hCoV-19/South Africa/NHLS-UCT-GP-5513/2020\|EPI_ISL | ···················· | ·A·················· | ···················· | ···················· | ···················· | ···················· | |

| | | | | | | | |
|---|---|---|---|---|---|---|---|
| W | 1894 hCoV-19/South Africa/NHLS-UCT-GP-5183/2020\|EPI_ISL | ···················· | ·A·················· | ···················· | ···················· | ···················· | ···················· |
| W | 1895 hCoV-19/South Africa/NHLS-UCT-GP-5720/2020\|EPI_ISL | ···················· | ···················· | ···················· | ···················· | ···················· | ···················· |
| W | 1896 hCoV-19/South Africa/NHLS-UCT-GS-0960/2020\|EPI_ISL | ···················· | ·A·················· | ···················· | ···················· | ···················· | ···················· |
| W | 1897 hCoV-19/South Africa/NHLS-UCT-GP-5790/2020\|EPI_ISL | ···················· | ···················· | ···················· | ···················· | ···················· | ···················· |
| W | 1898 hCoV-19/South Africa/NHLS-UCT-GS-1480/2020\|EPI_ISL | ···················· | ·A·················· | ···················· | ···················· | ···················· | ···················· |
| W | 1899 hCoV-19/South Africa/NHLS-UCT-GS-1763/2020\|EPI_ISL | ···················· | ·A·················· | ···················· | ···················· | ···················· | ···················· |
| W | 1900 hCoV-19/South Africa/NHLS-UCT-GP-5888/2020\|EPI_ISL | ···················· | ···················· | ···················· | ···················· | ···················· | ···················· |
| W | 1901 hCoV-19/South Africa/NHLS-UCT-GS-2980/2020\|EPI_ISL | ···················· | ···················· | ···················· | ···················· | ···················· | ···················· |
| W | 1902 hCoV-19/South Africa/NHLS-UCT-GS-1615/2020\|EPI_ISL | ···················· | ·A·················· | ···················· | ···················· | ···················· | ···················· |
| W | 1903 hCoV-19/South Africa/NHLS-UCT-GS-3168/2020\|EPI_ISL | ···················· | ···················· | ···················· | ···················· | ···················· | ···················· |
| W | 1904 hCoV-19/South Africa/NHLS-UCT-GS-2284/2020\|EPI_ISL | ···················· | ·A·················· | ···················· | ···················· | ···················· | ···················· |
| W | 1905 hCoV-19/South Africa/NHLS-UCT-GS-2911/2020\|EPI_ISL | ···················· | ···················· | ···················· | ···················· | ···················· | ···················· |
| W | 1907 hCoV-19/South Africa/NHLS-UCT-GP-5635/2020\|EPI_ISL | ···················· | ·A·················· | ···················· | ···················· | ···················· | ···················· |
| W | 1909 hCoV-19/South Africa/NHLS-UCT-GP-5332/2020\|EPI_ISL | ···················· | ·A·················· | ···················· | ···················· | ···················· | ···················· |
| W | 1910 hCoV-19/South Africa/NHLS-UCT-GS-1371/2020\|EPI_ISL | ···················· | ·A·················· | ···················· | ···················· | ···················· | ···················· |
| W | 1911 hCoV-19/South Africa/NHLS-UCT-GP-5349/2020\|EPI_ISL | ···················· | ·A·················· | ···················· | ···················· | ···················· | ···················· |
| W | 1912 hCoV-19/South Africa/NHLS-UCT-GS-2790/2020\|EPI_ISL | ···················· | ·A·················· | ···················· | ···················· | ···················· | ···················· |
| W | 1913 hCoV-19/South Africa/NHLS-UCT-GS-1530/2020\|EPI_ISL | ···················· | ·A·················· | ···················· | ···················· | ···················· | ···················· |
| W | 1915 hCoV-19/South Africa/NHLS-UCT-GS-0619/2020\|EPI_ISL | ···················· | ···················· | ···················· | ···················· | ···················· | ···················· |
| W | 1918 hCoV-19/South Africa/NHLS-UCT-GP-5164/2020\|EPI_ISL | ···················· | ·A·················· | ···················· | ···················· | ···················· | ···················· |
| W | 1919 hCoV-19/South Africa/NHLS-UCT-GP-5840/2020\|EPI_ISL | ···················· | ·A·················· | ···················· | ···················· | ···················· | ···················· |
| W | 1920 hCoV-19/South Africa/NHLS-UCT-GP-5890/2020\|EPI_ISL | ···················· | ···················· | ···················· | ···················· | ···················· | ···················· |
| W | 1921 hCoV-19/South Africa/NHLS-UCT-GS-2203/2020\|EPI_ISL | ···················· | ·A·················· | ···················· | ···················· | ···················· | ···················· |
| W | 1922 hCoV-19/South Africa/NHLS-UCT-GP-5597/2020\|EPI_ISL | ···················· | ···················· | ···················· | ···················· | ···················· | ···················· |
| W | 1923 hCoV-19/South Africa/NHLS-UCT-GS-1920/2020\|EPI_ISL | ···················· | ···················· | ···················· | ···················· | ···················· | ···················· |
| W | 1924 hCoV-19/South Africa/NHLS-UCT-GS-1808/2020\|EPI_ISL | ···················· | ·A·················· | ···················· | ···················· | ···················· | ···················· |
| W | 1925 hCoV-19/South Africa/NHLS-UCT-GS-2736/2020\|EPI_ISL | ···················· | ···················· | ···················· | ···················· | ···················· | ···················· |
| W | 1926 hCoV-19/South Africa/NHLS-UCT-GS-0654/2020\|EPI_ISL | ···················· | ···················· | ···················· | ···················· | ···················· | ···················· |
| W | 1927 hCoV-19/England/ALDP-BB3D28/2020\|EPI_ISL_701835\|20 | ···················· | ·A·················· | ···················· | ···················· | ···················· | ··A················· |
| W | 1928 hCoV-19/England/ALDP-BBAF58/2020\|EPI_ISL_701836\|20 | ···················· | ···················· | ···················· | ···················· | ···················· | ···················· |
| W | 1929 hCoV-19/England/ALDP-BB3818/2020\|EPI_ISL_701837\|20 | ···················· | ···················· | ···················· | ···················· | ···················· | ···················· |
| W | 1930 hCoV-19/England/ALDP-BB3D0A/2020\|EPI_ISL_701838\|20 | ···················· | ···················· | ···················· | ···················· | ·····T·············· | ···················· |
| W | 1931 hCoV-19/England/ALDP-BB9E6B/2020\|EPI_ISL_701840\|20 | ···················· | ···················· | ···················· | ···················· | ···················· | ···················· |
| W | 1932 hCoV-19/England/ALDP-BB3CEF/2020\|EPI_ISL_701841\|20 | ···················· | ···················· | ···················· | ···················· | ···················· | ···················· |
| W | 1933 hCoV-19/England/ALDP-BBB300/2020\|EPI_ISL_701844\|20 | ···················· | ···················· | ···················· | ···················· | ···················· | ···················· |
| W | 1934 hCoV-19/England/ALDP-BBB1F7/2020\|EPI_ISL_701845\|20 | ···················· | ···················· | ···················· | ···················· | ···················· | ···················· |
| W | 1935 hCoV-19/England/ALDP-BBA134/2020\|EPI_ISL_701846\|20 | ···················· | ···················· | ···················· | ···················· | ···················· | ···················· |
| W | 1936 hCoV-19/England/ALDP-BBAF1C/2020\|EPI_ISL_701847\|20 | ···················· | ···················· | ···················· | ···················· | ···················· | ···················· |
| W | 1937 hCoV-19/England/ALDP-BBA152/2020\|EPI_ISL_701849\|20 | ···················· | ···················· | ···················· | ···················· | ···················· | ···················· |
| W | 1938 hCoV-19/England/ALDP-BB3C2B/2020\|EPI_ISL_701850\|20 | ···················· | ···················· | ···················· | ···················· | ···················· | ···················· |
| W | 1939 hCoV-19/England/ALDP-BBA3B6/2020\|EPI_ISL_701852\|20 | ···················· | ···················· | ···················· | ···················· | ···················· | ···················· |
| W | 1940 hCoV-19/England/ALDP-BB3A30/2020\|EPI_ISL_701853\|20 | ···················· | ···················· | ···················· | ···················· | ···················· | ···················· |
| W | 1941 hCoV-19/England/ALDP-BB9EA7/2020\|EPI_ISL_701854\|20 | ···················· | ···················· | ···················· | ···················· | ···················· | ···················· |
| W | 1942 hCoV-19/England/ALDP-BB9F1D/2020\|EPI_ISL_701855\|20 | ···················· | ···················· | ···················· | ···················· | ···················· | ···················· |
| W | 1943 hCoV-19/England/ALDP-BBB1CA/2020\|EPI_ISL_701856\|20 | ···················· | ···················· | ···················· | ···················· | ···················· | ···················· |
| W | 1944 hCoV-19/England/ALDP-BB4FE5/2020\|EPI_ISL_701874\|20 | ···················· | ···················· | ···················· | ···················· | ···················· | ···················· |
| W | 1945 hCoV-19/England/ALDP-BB3D19/2020\|EPI_ISL_701875\|20 | ···················· | ···················· | ···················· | ···················· | ···················· | ···················· |
| W | 1946 hCoV-19/England/ALDP-BB53F7/2020\|EPI_ISL_701877\|20 | ···················· | ···················· | ···················· | ···················· | ···················· | ···················· |
| W | 1947 hCoV-19/England/ALDP-BB3854/2020\|EPI_ISL_701878\|20 | ···················· | ···················· | ···················· | ···················· | ···················· | ···················· |
| W | 1948 hCoV-19/England/ALDP-BBB388/2020\|EPI_ISL_701879\|20 | ···················· | ···················· | ···················· | ···················· | ···················· | ···················· |
| W | 1949 hCoV-19/England/ALDP-BBA028/2020\|EPI_ISL_701880\|20 | ···················· | ···················· | ···················· | ···················· | ···················· | ···················· |
| W | 1950 hCoV-19/England/ALDP-BBA0EC/2020\|EPI_ISL_701882\|20 | ···················· | ···················· | ···················· | ···················· | ···················· | ···················· |
| W | 1951 hCoV-19/England/ALDP-BB9EF2/2020\|EPI_ISL_701883\|20 | ···················· | ···················· | ···················· | ···················· | ···················· | ···················· |
| W | 1952 hCoV-19/England/ALDP-BB503C/2020\|EPI_ISL_701884\|20 | ···················· | ···················· | ···················· | ···················· | ···················· | ···················· |
| W | 1953 hCoV-19/England/ALDP-BBB115/2020\|EPI_ISL_701885\|20 | ···················· | ···················· | ···················· | ···················· | ···················· | ···················· |
| W | 1954 hCoV-19/England/ALDP-BB9F68/2020\|EPI_ISL_701887\|20 | ···················· | ···················· | ···················· | ···················· | ···················· | ···················· |
| W | 1955 hCoV-19/England/ALDP-BB5209/2020\|EPI_ISL_701888\|20 | ···················· | ···················· | ···················· | ···················· | ···················· | ···················· |
| W | 1956 hCoV-19/England/ALDP-BB51A2/2020\|EPI_ISL_701889\|20 | ···················· | ···················· | ···················· | ···················· | ···················· | ···················· |
| W | 1957 hCoV-19/England/ALDP-BB9FE0/2020\|EPI_ISL_701890\|20 | ···················· | ···················· | ···················· | ···················· | ···················· | ···················· |
| W | 1958 hCoV-19/England/ALDP-BBA84E/2020\|EPI_ISL_701891\|20 | ···················· | ···················· | ···················· | ···················· | ·····T·············· | ···················· |
| W | 1959 hCoV-19/England/ALDP-BBA161/2020\|EPI_ISL_701892\|20 | ···················· | ···················· | ···················· | ···················· | ···················· | ···················· |
| W | 1960 hCoV-19/England/ALDP-BBA0CE/2020\|EPI_ISL_701893\|20 | ···················· | ···················· | ···················· | ···················· | ···················· | ···················· |
| W | 1961 hCoV-19/England/ALDP-BBAFEF/2020\|EPI_ISL_701894\|20 | ···················· | ···················· | ···················· | ···················· | ···················· | ···················· |
| W | 1962 hCoV-19/England/ALDP-BBB027/2020\|EPI_ISL_701895\|20 | ···················· | ···················· | ···················· | ···················· | ···················· | ···················· |
| W | 1963 hCoV-19/England/ALDP-BB545E/2020\|EPI_ISL_701896\|20 | ···················· | ···················· | ···················· | ···················· | ···················· | ···················· |
| W | 1964 hCoV-19/England/ALDP-BB502D/2020\|EPI_ISL_701897\|20 | ···················· | ···················· | ···················· | ···················· | ···················· | ···················· |
| W | 1965 hCoV-19/England/ALDP-BBA77E/2020\|EPI_ISL_701898\|20 | ···················· | ···················· | ···················· | ···················· | ···················· | ···················· |
| W | 1966 hCoV-19/England/ALDP-BB3915/2020\|EPI_ISL_701899\|20 | ···················· | ···················· | ···················· | ···················· | ···················· | ··A················· |
| W | 1967 hCoV-19/England/QEUH-BC3DAD/2020\|EPI_ISL_704103\|20 | ···················· | ···················· | ···················· | ···················· | ···················· | ···················· |
| W | 1968 hCoV-19/England/QEUH-BCB419/2020\|EPI_ISL_704104\|20 | ···················· | ···················· | ···················· | ···················· | ···················· | ···················· |
| W | 1969 hCoV-19/England/QEUH-BCB83B/2020\|EPI_ISL_704106\|20 | ···················· | ···················· | ···················· | ···················· | ···················· | ···················· |
| W | 1970 hCoV-19/England/QEUH-BCC73D/2020\|EPI_ISL_704107\|20 | ···················· | ···················· | ···················· | ···················· | ···················· | ···················· |
| W | 1971 hCoV-19/England/QEUH-BC3763/2020\|EPI_ISL_704109\|20 | ···················· | ···················· | ···················· | ···················· | ···················· | ···················· |
| W | 1972 hCoV-19/Wales/PHWC-491CC0/2020\|EPI_ISL_705420\|2020 | ···················· | ···················· | ···················· | ···················· | ···················· | ···················· |
| W | 1973 hCoV-19/England/SHEF-10C7600/2020\|EPI_ISL_705421\|2 | ···················· | ···················· | ···················· | ···················· | ···················· | ···················· |
| W | 1974 hCoV-19/England/CAMC-BBCCAA/2020\|EPI_ISL_705422\|20 | ···················· | ···················· | ···················· | ···················· | ···················· | ···················· |
| W | 1975 hCoV-19/Wales/LIVE-DB976E/2020\|EPI_ISL_705423\|2020 | ···················· | ···················· | ···················· | ···················· | ···················· | ···················· |
| W | 1976 hCoV-19/England/LIVE-DB993B/2020\|EPI_ISL_705424\|20 | ···················· | ···················· | ···················· | ···················· | ···················· | ···················· |
| W | 1977 hCoV-19/England/LOND-12D4DEB/2020\|EPI_ISL_705425\|2 | ···················· | ···················· | ···················· | ···················· | ···················· | ···················· |
| W | 1978 hCoV-19/England/MILK-B3B371/2020\|EPI_ISL_705426\|20 | ···················· | ···················· | ···················· | ···················· | ···················· | ···················· |

| | | | | | | | | | |
|---|---|---|---|---|---|---|---|---|---|
| W | 1979 hCoV-19/England/MILK-B3B942/2020\|EPI_ISL_705427\|20 | ···················· | ···················· | ···················· | ···················· | ···················· | ···················· | | |
| W | 1980 hCoV-19/England/MILK-B3C228/2020\|EPI_ISL_705428\|20 | ···················· | ···················· | ···················· | ···················· | ···················· | ···················· | | |
| W | 1981 hCoV-19/England/MILK-B3C914/2020\|EPI_ISL_705429\|20 | ···················· | ···················· | ···················· | ···················· | ···················· | ···················· | | |
| W | 1982 hCoV-19/England/MILK-B3D8CB/2020\|EPI_ISL_705430\|20 | ···················· | ···················· | ···················· | ···················· | ···················· | ···················· | | |
| W | 1983 hCoV-19/England/NORW-F750D/2020\|EPI_ISL_705431\|202 | ···················· | ·A·················· | ···················· | ···················· | ···················· | ··A················· | | |
| W | 1984 hCoV-19/England/NORW-F8AFE/2020\|EPI_ISL_705432\|202 | ···················· | ···················· | ···················· | ···················· | ···················· | ···················· | | |
| W | 1986 hCoV-19/Wales/PHWC-490BA6/2020\|EPI_ISL_705434\|2020 | ···················· | ···················· | ···················· | ···················· | ···················· | ···················· | | |
| W | 1987 hCoV-19/Wales/PHWC-490F04/2020\|EPI_ISL_705435\|2020 | ···················· | ···················· | ···················· | ···················· | NNN················· | ···················· | | 1 |
| W | 1988 hCoV-19/Wales/PHWC-4917B0/2020\|EPI_ISL_705436\|2020 | ···················· | ···················· | ···················· | ···················· | ···················· | ···················· | | |
| W | 1989 hCoV-19/Wales/PHWC-491808/2020\|EPI_ISL_705437\|2020 | ···················· | ···················· | ···················· | ···················· | ···················· | ···················· | | |
| W | 1990 hCoV-19/Wales/PHWC-491844/2020\|EPI_ISL_705438\|2020 | ···················· | ···················· | ···················· | ···················· | ···················· | ···················· | | |
| W | 1991 hCoV-19/Wales/PHWC-491DDC/2020\|EPI_ISL_705439\|2020 | ···················· | ···················· | ···················· | ···················· | ···················· | ···················· | | |
| W | 1992 hCoV-19/Wales/PHWC-491862/2020\|EPI_ISL_706700\|2020 | ···················· | ···················· | ···················· | ···················· | ···················· | ···················· | | |
| W | 1993 hCoV-19/Wales/PHWC-491871/2020\|EPI_ISL_706701\|2020 | ···················· | ···················· | ···················· | ···················· | ···················· | ···················· | | |
| W | 1994 hCoV-19/Wales/PHWC-491880/2020\|EPI_ISL_706702\|2020 | ···················· | ···················· | ···················· | ···················· | ···················· | ···················· | | |
| W | 1995 hCoV-19/Wales/PHWC-49189F/2020\|EPI_ISL_706703\|2020 | ···················· | ···················· | ···················· | ···················· | ···················· | ···················· | | |
| W | 1997 hCoV-19/Wales/PHWC-4918BD/2020\|EPI_ISL_706705\|2020 | ···················· | ···················· | ···················· | ···················· | ···················· | ···················· | | |
| W | 2002 hCoV-19/Wales/PHWC-491905/2020\|EPI_ISL_706710\|2020 | ···················· | ···················· | ···················· | ···················· | ···················· | ···················· | | |
| W | 2003 hCoV-19/Wales/PHWC-491914/2020\|EPI_ISL_706711\|2020 | ···················· | ···················· | ···················· | ···················· | ···················· | ···················· | | |
| W | 2004 hCoV-19/Wales/PHWC-491923/2020\|EPI_ISL_706712\|2020 | ···················· | ···················· | ···················· | ···················· | ···················· | ···················· | | |
| W | 2006 hCoV-19/Wales/PHWC-491941/2020\|EPI_ISL_706714\|2020 | ···················· | ···················· | ···················· | ···················· | ···················· | ···················· | | |
| W | 2007 hCoV-19/Wales/PHWC-491950/2020\|EPI_ISL_706715\|2020 | ···················· | ···················· | ···················· | ···················· | ···················· | ···················· | | |
| W | 2008 hCoV-19/Wales/PHWC-49196F/2020\|EPI_ISL_706716\|2020 | ···················· | ···················· | ···················· | ···················· | ···················· | ···················· | | |
| W | 2011 hCoV-19/Wales/PHWC-49199C/2020\|EPI_ISL_706719\|2020 | ···················· | ···················· | ···················· | ···················· | ···················· | ···················· | | |
| W | 2013 hCoV-19/Wales/PHWC-4919BA/2020\|EPI_ISL_706721\|2020 | ···················· | ···················· | ···················· | ···················· | ···················· | ···················· | | |
| W | 2014 hCoV-19/Wales/PHWC-4919C9/2020\|EPI_ISL_706722\|2020 | ···················· | ···················· | ···················· | ···················· | ···················· | ···················· | | |
| W | 2018 hCoV-19/Wales/PHWC-491A02/2020\|EPI_ISL_706726\|2020 | ···················· | ···················· | ···················· | ···················· | ···················· | ···················· | | |
| W | 2020 hCoV-19/Wales/PHWC-491A20/2020\|EPI_ISL_706728\|2020 | ···················· | ···················· | ···················· | ···················· | ···················· | ···················· | | |
| W | 2022 hCoV-19/Wales/PHWC-491A4E/2020\|EPI_ISL_706730\|2020 | ···················· | ···················· | ···················· | ···················· | ···················· | ···················· | | |
| W | 2023 hCoV-19/Wales/PHWC-491A5D/2020\|EPI_ISL_706731\|2020 | ···················· | ···················· | ···················· | ···················· | NNN················· | ···················· | | 1 |
| W | 2024 hCoV-19/Wales/PHWC-491A6C/2020\|EPI_ISL_706732\|2020 | ···················· | ···················· | ···················· | ···················· | ···················· | ···················· | | |
| W | 2026 hCoV-19/Wales/PHWC-491A8A/2020\|EPI_ISL_706734\|2020 | ···················· | ···················· | ···················· | ···················· | ···················· | ···················· | | |
| W | 2029 hCoV-19/Wales/PHWC-491B87/2020\|EPI_ISL_706750\|2020 | ···················· | ···················· | ···················· | ···················· | ···················· | ···················· | | |
| W | 2031 hCoV-19/Wales/PHWC-491BA5/2020\|EPI_ISL_706752\|2020 | ···················· | ···················· | ···················· | ···················· | ··················NN | ···················· | 1 | |
| W | 2032 hCoV-19/Wales/PHWC-491BB4/2020\|EPI_ISL_706753\|2020 | ···················· | ···················· | ···················· | ···················· | ···················· | ···················· | | |
| W | 2033 hCoV-19/Wales/PHWC-491BC3/2020\|EPI_ISL_706754\|2020 | ···················· | ···················· | ···················· | ···················· | ···················· | ···················· | | |
| W | 2035 hCoV-19/Wales/PHWC-491BE1/2020\|EPI_ISL_706756\|2020 | ···················· | ···················· | ···················· | ···················· | ···················· | ···················· | | |
| W | 2036 hCoV-19/Wales/PHWC-491BF0/2020\|EPI_ISL_706757\|2020 | ···················· | ···················· | ···················· | ···················· | ···················· | ···················· | | |
| W | 2037 hCoV-19/Wales/PHWC-491C1B/2020\|EPI_ISL_706758\|2020 | ···················· | ···················· | ···················· | ···················· | ···················· | ···················· | | |
| W | 2039 hCoV-19/Wales/PHWC-491C39/2020\|EPI_ISL_706760\|2020 | ···················· | ···················· | ···················· | ···················· | ···················· | ···················· | | |
| W | 2040 hCoV-19/Wales/PHWC-491C48/2020\|EPI_ISL_706761\|2020 | ···················· | ···················· | ···················· | ···················· | ···················· | ···················· | | |
| W | 2042 hCoV-19/Wales/PHWC-491C84/2020\|EPI_ISL_706763\|2020 | ···················· | ···················· | ···················· | ···················· | ···················· | ···················· | | |
| W | 2043 hCoV-19/Wales/PHWC-491C93/2020\|EPI_ISL_706764\|2020 | ···················· | ···················· | ···················· | ···················· | ···················· | ···················· | | |
| W | 2044 hCoV-19/Wales/PHWC-491CA2/2020\|EPI_ISL_706765\|2020 | ···················· | ···················· | ···················· | ···················· | ···················· | ···················· | | |
| W | 2045 hCoV-19/Wales/PHWC-491CB1/2020\|EPI_ISL_706766\|2020 | ···················· | ···················· | ···················· | ···················· | ···················· | ···················· | | |
| W | 2046 hCoV-19/Wales/PHWC-491CDF/2020\|EPI_ISL_706767\|2020 | ···················· | ···················· | ···················· | ···················· | ···················· | NN·················· | | |
| W | 2047 hCoV-19/Wales/PHWC-491CEE/2020\|EPI_ISL_706768\|2020 | ···················· | ···················· | ···················· | ···················· | ···················· | ···················· | | |
| W | 2048 hCoV-19/Wales/PHWC-491CFD/2020\|EPI_ISL_706769\|2020 | ···················· | ···················· | ···················· | ···················· | ···················· | ···················· | | |
| W | 2049 hCoV-19/USA/AZ-TG280866/2020\|EPI_ISL_694200\|2020-0 | ···················· | ···················· | ···················· | ···················· | ···················· | ···················· | | |
| W | 2050 hCoV-19/USA/AZ-TG280874/2020\|EPI_ISL_694205\|2020-0 | ···················· | ···················· | ···················· | ···················· | ···················· | ···················· | | |
| W | 2051 hCoV-19/USA/AZ-TG280876/2020\|EPI_ISL_694207\|2020-0 | ···················· | ···················· | ···················· | ···················· | ···················· | ···················· | | |
| W | 2052 hCoV-19/USA/AZ-TG280877/2020\|EPI_ISL_694208\|2020-0 | ···················· | ···················· | ···················· | ···················· | ···················· | ···················· | | |
| W | 2053 hCoV-19/USA/AZ-TG280881/2020\|EPI_ISL_694210\|2020-0 | ···················· | ···················· | ···················· | ···················· | ···················· | ···················· | | |
| W | 2054 hCoV-19/USA/AZ-TG280882/2020\|EPI_ISL_694211\|2020-0 | ···················· | ···················· | ···················· | ···················· | ···················· | ···················· | | |
| W | 2055 hCoV-19/USA/AZ-TG280883/2020\|EPI_ISL_694212\|2020-0 | ···················· | ···················· | ···················· | ···················· | ···················· | ···················· | | |
| W | 2056 hCoV-19/USA/AZ-TG280893/2020\|EPI_ISL_694218\|2020-0 | ···················· | ···················· | ···················· | ···················· | ···················· | ···················· | | |
| W | 2057 hCoV-19/USA/AZ-TG281238/2020\|EPI_ISL_694222\|2020-0 | ···················· | ···················· | ···················· | ···················· | ···················· | ···················· | | |
| W | 2058 hCoV-19/USA/AZ-TG281380/2020\|EPI_ISL_694223\|2020-0 | ···················· | ···················· | ···················· | ···················· | ···················· | ···················· | | |
| W | 2059 hCoV-19/USA/AZ-TG281576/2020\|EPI_ISL_694224\|2020-0 | ···················· | ···················· | ···················· | ···················· | ···················· | ···················· | | |
| W | 2060 hCoV-19/USA/AZ-TG283957/2020\|EPI_ISL_694228\|2020-0 | ···················· | ···················· | ···················· | ···················· | ···················· | ···················· | | |
| W | 2061 hCoV-19/USA/AZ-TG287820/2020\|EPI_ISL_694233\|2020-0 | ···················· | ···················· | ···················· | ···················· | ···················· | ···················· | | |
| W | 2062 hCoV-19/USA/AZ-TG290723/2020\|EPI_ISL_694239\|2020-0 | ···················· | ···················· | ···················· | ···················· | ···················· | ···················· | | |
| W | 2063 hCoV-19/USA/AZ-TG295775/2020\|EPI_ISL_694242\|2020-0 | ···················· | ···················· | ···················· | ···················· | ···················· | ···················· | | |
| W | 2064 hCoV-19/USA/AZ-TG296708/2020\|EPI_ISL_694244\|2020-0 | ···················· | ···················· | ···················· | ···················· | ···················· | ···················· | | |
| W | 2065 hCoV-19/USA/AZ-TG299365/2020\|EPI_ISL_694247\|2020-0 | ···················· | ···················· | ···················· | ···················· | ···················· | ···················· | | |
| W | 2066 hCoV-19/USA/AZ-TG305701/2020\|EPI_ISL_694249\|2020-0 | ···················· | ···················· | ···················· | ···················· | ···················· | ···················· | | |
| W | 2067 hCoV-19/USA/AZ-TG305703/2020\|EPI_ISL_694250\|2020-0 | ···················· | ···················· | ···················· | ···················· | ···················· | ···················· | | |
| W | 2068 hCoV-19/USA/AZ-TG305707/2020\|EPI_ISL_694252\|2020-0 | ···················· | ···················· | ···················· | ···················· | ···················· | ···················· | | |
| W | 2069 hCoV-19/USA/AZ-TG305713/2020\|EPI_ISL_694254\|2020-0 | ···················· | ···················· | ···················· | ···················· | ···················· | ···················· | | |
| W | 2070 hCoV-19/USA/AZ-TG305718/2020\|EPI_ISL_694255\|2020-0 | ···················· | ···················· | ···················· | ···················· | ···················· | ···················· | | |
| W | 2071 hCoV-19/USA/AZ-TG305720/2020\|EPI_ISL_694256\|2020-0 | ···················· | ···················· | ···················· | ···················· | ···················· | ···················· | | |
| W | 2072 hCoV-19/USA/AZ-TG305728/2020\|EPI_ISL_694258\|2020-0 | ···················· | ···················· | ···················· | ···················· | ···················· | ···················· | | |
| W | 2073 hCoV-19/USA/AZ-TG305764/2020\|EPI_ISL_694269\|2020-0 | ···················· | ···················· | ···················· | ···················· | ···················· | ···················· | | |
| W | 2074 hCoV-19/USA/AZ-TG305766/2020\|EPI_ISL_694270\|2020-0 | ···················· | ···················· | ···················· | ···················· | ···················· | ···················· | | |
| W | 2075 hCoV-19/USA/AZ-TG305770/2020\|EPI_ISL_694272\|2020-0 | ···················· | ···················· | ···················· | ···················· | ···················· | ···················· | | |
| W | 2076 hCoV-19/USA/AZ-TG305787/2020\|EPI_ISL_694276\|2020-0 | ···················· | ···················· | ···················· | ···················· | ···················· | ···················· | | |
| W | 2077 hCoV-19/USA/AZ-TG305789/2020\|EPI_ISL_694277\|2020-0 | ···················· | ···················· | ···················· | ···················· | ···················· | ···················· | | |
| W | 2078 hCoV-19/USA/AZ-TG305800/2020\|EPI_ISL_694280\|2020-0 | ···················· | ···················· | ···················· | ···················· | ···················· | ···················· | | |
| W | 2079 hCoV-19/USA/AZ-TG305865/2020\|EPI_ISL_694283\|2020-0 | ···················· | ···················· | ···················· | ···················· | ···················· | ···················· | | |
| W | 2080 hCoV-19/USA/AZ-TG305867/2020\|EPI_ISL_694284\|2020-0 | ···················· | ···················· | ···················· | ···················· | ···················· | ···················· | | |

| | | | | | | | |
|---|---|---|---|---|---|---|---|
| W | 2081 hCoV-19/USA/AZ-TG305869/2020\|EPI_ISL_694285\|2020-0 | .................... | .................... | ..................... | .................... | .................... | .................... |
| W | 2082 hCoV-19/USA/AZ-TG305893/2020\|EPI_ISL_694289\|2020-0 | .................... | .................... | ..................... | .................... | .................... | .................... |
| W | 2083 hCoV-19/USA/AZ-TG305897/2020\|EPI_ISL_694290\|2020-0 | .................... | .................... | ..................... | .................... | .................... | .................... |
| W | 2084 hCoV-19/USA/AZ-TG305899/2020\|EPI_ISL_694291\|2020-0 | .................... | .................... | ..................... | .................... | .................... | .................... |
| W | 2085 hCoV-19/USA/AZ-TG305901/2020\|EPI_ISL_694292\|2020-0 | .................... | .................... | ..................... | .................... | .................... | .................... |
| W | 2086 hCoV-19/USA/AZ-TG305908/2020\|EPI_ISL_694294\|2020-0 | .................... | .................... | ..................... | .................... | .................... | .................... |
| W | 2087 hCoV-19/USA/AZ-TG305914/2020\|EPI_ISL_694297\|2020-0 | .................... | .................... | ..................... | .................... | .................... | .................... |
| W | 2088 hCoV-19/USA/AZ-TG305956/2020\|EPI_ISL_694298\|2020-0 | .................... | .................... | ..................... | .................... | .................... | .................... |
| W | 2089 hCoV-19/USA/AZ-TG305958/2020\|EPI_ISL_694299\|2020-0 | .................... | .................... | ..................... | .................... | .................... | .................... |
| W | 2090 hCoV-19/USA/AZ-TG547354/2020\|EPI_ISL_695500\|2020-0 | .................... | .A.................. | ..................... | .................... | .................... | .................... |
| W | 2091 hCoV-19/USA/AZ-TG547355/2020\|EPI_ISL_695501\|2020-0 | .................... | .A.................. | ..................... | .................... | .................... | .................... |
| W | 2092 hCoV-19/USA/AZ-TG547357/2020\|EPI_ISL_695502\|2020-0 | .................... | .A.................. | ..................... | .................... | .................... | .................... |
| W | 2093 hCoV-19/USA/AZ-TG547363/2020\|EPI_ISL_695506\|2020-0 | .................... | .A.................. | ..................... | .................... | .................... | .................... |
| W | 2094 hCoV-19/USA/AZ-TG547382/2020\|EPI_ISL_695514\|2020-0 | .................... | .A.................. | ..................... | .................... | .................... | .................... |
| W | 2095 hCoV-19/USA/AZ-TG547386/2020\|EPI_ISL_695515\|2020-0 | .................... | .A.................. | ..................... | .................... | .................... | .................... |
| W | 2096 hCoV-19/USA/AZ-TG547388/2020\|EPI_ISL_695516\|2020-0 | .................... | .A.................. | ..................... | .................... | .................... | .................... |
| W | 2097 hCoV-19/USA/AZ-TG547409/2020\|EPI_ISL_695521\|2020-0 | .................... | .A.................. | ..................... | .................... | .................... | .................... |
| W | 2098 hCoV-19/USA/AZ-TG547912/2020\|EPI_ISL_695522\|2020-0 | .................... | .................... | ..................... | .................... | .................... | .................... |
| W | 2099 hCoV-19/USA/AZ-TG548700/2020\|EPI_ISL_695526\|2020-0 | .................... | .A.................. | ..................... | .................... | .................... | .................... |
| W | 2100 hCoV-19/USA/AZ-TG550900/2020\|EPI_ISL_695532\|2020-0 | .................... | .................... | ..................... | .................... | .................... | .................... |
| W | 2101 hCoV-19/USA/AZ-TG551036/2020\|EPI_ISL_695533\|2020-0 | .................... | .................... | ..................... | .................... | .................... | .................... |
| W | 2102 hCoV-19/USA/AZ-TG552537/2020\|EPI_ISL_695535\|2020-0 | .................... | .A.................. | ..................... | .................... | .................... | .................... |
| W | 2103 hCoV-19/USA/AZ-TG553089/2020\|EPI_ISL_695536\|2020-0 | .................... | .A.................. | ..................... | .................... | .................... | .................... |
| W | 2104 hCoV-19/USA/AZ-TG553104/2020\|EPI_ISL_695537\|2020-0 | .................... | .................... | ..................... | .................... | .................... | .................... |
| W | 2105 hCoV-19/USA/AZ-TG554086/2020\|EPI_ISL_695538\|2020-0 | .................... | .................... | ..................... | .................... | .................... | .................... |
| W | 2106 hCoV-19/USA/AZ-TG554161/2020\|EPI_ISL_695539\|2020-0 | .................... | .................... | ..................... | .................... | .................... | .................... |
| W | 2107 hCoV-19/USA/AZ-TG555863/2020\|EPI_ISL_695543\|2020-0 | .................... | .................... | ..................... | .................... | .................... | .................... |
| W | 2108 hCoV-19/USA/AZ-TG557127/2020\|EPI_ISL_695544\|2020-0 | .................... | .................... | ..................... | .................... | .................... | .................... |
| W | 2109 hCoV-19/USA/AZ-TG558317/2020\|EPI_ISL_695545\|2020-0 | .................... | .A.................. | ..................... | .................... | .................... | .................... |
| W | 2110 hCoV-19/USA/AZ-TG558837/2020\|EPI_ISL_695547\|2020-0 | .................... | .................... | ..................... | .................... | .................... | .................... |
| W | 2111 hCoV-19/USA/AZ-TG558838/2020\|EPI_ISL_695548\|2020-0 | .................... | .................... | ..................... | .................... | .................... | .................... |
| W | 2112 hCoV-19/USA/AZ-TG561151/2020\|EPI_ISL_695550\|2020-0 | .................... | .A.................. | ..................... | .................... | .................... | .................... |
| W | 2113 hCoV-19/USA/AZ-TG563168/2020\|EPI_ISL_695553\|2020-0 | .................... | .................... | ..................... | .................... | .................... | .................... |
| W | 2114 hCoV-19/USA/AZ-TG565575/2020\|EPI_ISL_695555\|2020-0 | .................... | .A.................. | ..................... | .................... | .................... | .................... |
| W | 2115 hCoV-19/USA/AZ-TG565579/2020\|EPI_ISL_695556\|2020-0 | .................... | .................... | ..................... | .................... | .................... | .................... |
| W | 2116 hCoV-19/USA/AZ-TG565817/2020\|EPI_ISL_695557\|2020-0 | .................... | .................... | ..................... | .................... | .................... | .................... |
| W | 2117 hCoV-19/USA/AZ-TG565842/2020\|EPI_ISL_695558\|2020-0 | .................... | .A.................. | ..................... | .................... | .................... | .................... |
| W | 2118 hCoV-19/USA/AZ-TG566378/2020\|EPI_ISL_695561\|2020-0 | .................... | .................... | ..................... | .................... | .................... | .................... |
| W | 2119 hCoV-19/USA/AZ-TG569465/2020\|EPI_ISL_695568\|2020-0 | .................... | .................... | ..................... | .................... | .................... | .................... |
| W | 2120 hCoV-19/USA/AZ-TG569471/2020\|EPI_ISL_695571\|2020-0 | .................... | .................... | ..................... | .................... | .................... | .................... |
| W | 2121 hCoV-19/USA/AZ-TG569484/2020\|EPI_ISL_695578\|2020-0 | .................... | .A.................. | ..................... | .................... | .................... | .................... |
| W | 2122 hCoV-19/USA/AZ-TG569487/2020\|EPI_ISL_695579\|2020-0 | .................... | .A.................. | ..................... | .................... | .................... | .................... |
| W | 2123 hCoV-19/USA/AZ-TG569502/2020\|EPI_ISL_695586\|2020-0 | .................... | .................... | ..................... | .................... | .................... | .................... |
| W | 2124 hCoV-19/USA/AZ-TG569504/2020\|EPI_ISL_695588\|2020-0 | .................... | .................... | ..................... | .................... | .................... | .................... |
| W | 2125 hCoV-19/USA/AZ-TG569513/2020\|EPI_ISL_695595\|2020-0 | .................... | .................... | ..................... | .................... | .................... | .................... |
| W | 2126 hCoV-19/USA/AZ-TG569518/2020\|EPI_ISL_695598\|2020-0 | .................... | .................... | ..................... | .................... | .................... | .................... |
| W | 2127 hCoV-19/USA/AZ-TG664299/2020\|EPI_ISL_696093\|2020-0 | .................... | .................... | ..................... | .................... | .................... | .................... |
| W | 2128 hCoV-19/USA/AZ-TG664300/2020\|EPI_ISL_696094\|2020-0 | .................... | .................... | ..................... | .................... | .................... | .................... |
| W | 2129 hCoV-19/United Arab Emirates/0869/2020\|EPI_ISL_699 | .................... | .................... | ..................... | .................... | .................... | .................... |
| W | 2130 hCoV-19/United Arab Emirates/0870/2020\|EPI_ISL_699 | .................... | .................... | ..................... | .................... | .................... | .................... |
| W | 2131 hCoV-19/United Arab Emirates/0871/2020\|EPI_ISL_699 | .................... | .................... | ..................... | .................... | .................... | .................... |
| W | 2132 hCoV-19/United Arab Emirates/0872/2020\|EPI_ISL_699 | .................... | .................... | ..................... | .................... | .................... | .................... |
| W | 2133 hCoV-19/United Arab Emirates/0873/2020\|EPI_ISL_699 | .................... | .................... | ..................... | .................... | .................... | .................... |
| W | 2134 hCoV-19/United Arab Emirates/0874/2020\|EPI_ISL_699 | .................... | .................... | ..................... | .................... | .................... | .................... |
| W | 2135 hCoV-19/United Arab Emirates/0875/2020\|EPI_ISL_699 | .................... | .................... | ..................... | .................... | .................... | .................... |
| W | 2136 hCoV-19/United Arab Emirates/0876/2020\|EPI_ISL_699 | .................... | .................... | ..................... | .................... | .................... | .................... |
| W | 2137 hCoV-19/United Arab Emirates/0878/2020\|EPI_ISL_699 | .................... | .................... | ..................... | .................... | .................... | .................... |
| W | 2138 hCoV-19/United Arab Emirates/0879/2020\|EPI_ISL_699 | .................... | .................... | ..................... | .................... | .................... | .................... |
| W | 2139 hCoV-19/United Arab Emirates/0880/2020\|EPI_ISL_699 | .................... | .................... | ..................... | .................... | .................... | .................... |
| W | 2140 hCoV-19/United Arab Emirates/0881/2020\|EPI_ISL_699 | .................... | .................... | ..................... | .................... | .................... | .................... |
| W | 2141 hCoV-19/United Arab Emirates/0883/2020\|EPI_ISL_699 | .................... | .................... | ..................... | .................... | .................... | .................... |
| W | 2142 hCoV-19/United Arab Emirates/0884/2020\|EPI_ISL_699 | .................... | .................... | ..................... | .................... | .................... | .................... |
| W | 2143 hCoV-19/United Arab Emirates/0885/2020\|EPI_ISL_699 | .................... | .................... | ..................... | .................... | .................... | .................... |
| W | 2144 hCoV-19/United Arab Emirates/0886/2020\|EPI_ISL_699 | .................... | .................... | ..................... | .................... | .................... | .................... |
| W | 2145 hCoV-19/United Arab Emirates/0887/2020\|EPI_ISL_699 | .................... | .................... | ..................... | .................... | .................... | .................... |
| W | 2146 hCoV-19/United Arab Emirates/0888/2020\|EPI_ISL_699 | .................... | .................... | ..................... | .................... | .................... | .................... |
| W | 2147 hCoV-19/United Arab Emirates/0889/2020\|EPI_ISL_699 | .................... | .................... | ..................... | .................... | .................... | .................... |
| W | 2148 hCoV-19/United Arab Emirates/0890/2020\|EPI_ISL_699 | .................... | .................... | ..................... | .................... | .................... | .................... |
| W | 2149 hCoV-19/United Arab Emirates/0891/2020\|EPI_ISL_699 | .................... | .................... | ..................... | .................... | .................... | .................... |
| W | 2150 hCoV-19/United Arab Emirates/0954/2020\|EPI_ISL_699 | .................... | .................... | ..................... | .................... | .................... | .................... |
| W | 2151 hCoV-19/United Arab Emirates/0955/2020\|EPI_ISL_699 | .................... | .................... | ..................... | .................... | .................... | .................... |
| W | 2152 hCoV-19/United Arab Emirates/0956/2020\|EPI_ISL_699 | .................... | .................... | ..................... | .................... | .................... | .................... |
| W | 2153 hCoV-19/United Arab Emirates/0957/2020\|EPI_ISL_699 | .................... | .................... | ..................... | .................... | .................... | .................... |
| W | 2154 hCoV-19/United Arab Emirates/0958/2020\|EPI_ISL_699 | .................... | .................... | ..................... | .................... | .................... | .................... |
| W | 2155 hCoV-19/United Arab Emirates/0959/2020\|EPI_ISL_699 | .................... | .................... | ..................... | .................... | .................... | .................... |
| W | 2156 hCoV-19/United Arab Emirates/0960/2020\|EPI_ISL_699 | .................... | .................... | ..................... | .................... | .................... | .................... |
| W | 2157 hCoV-19/United Arab Emirates/0961/2020\|EPI_ISL_699 | .................... | .A.................. | ..................... | .................... | .................... | .................... |
| W | 2158 hCoV-19/United Arab Emirates/0962/2020\|EPI_ISL_699 | .................... | .................... | ..................... | .................... | .................... | .................... |
| W | 2159 hCoV-19/United Arab Emirates/0963/2020\|EPI_ISL_699 | .................... | .................... | ..................... | .................... | .................... | .................... |
| W | 2160 hCoV-19/England/ALDP-BB4A11/2020\|EPI_ISL_702400\|20 | .................... | .................... | ..................... | .................... | .................... | .................... |

| | | | | | | | | |
|---|---|---|---|---|---|---|---|---|
| W | 2161 hCoV-19/England/ALDP-BB934E/2020\|EPI_ISL_702401\|20 | ···················· | ···················· | ···················· | ···················· | ···················· | ···················· | |
| W | 2163 hCoV-19/Wales/PHWC-4908BE/2020\|EPI_ISL_705413\|2020 | ···················· | ···················· | ···················· | ···················· | ···················· | ···················· | |
| W | 2164 hCoV-19/England/CAMB-1B90F6/2020\|EPI_ISL_705415\|20 | ···················· | ···················· | ···················· | ···················· | ···················· | ···················· | |
| W | 2165 hCoV-19/Scotland/EDB769/2020\|EPI_ISL_705416\|2020-0 | ···················· | ···················· | ···················· | ···················· | ···················· | ···················· | |
| W | 2166 hCoV-19/England/MILK-A91969/2020\|EPI_ISL_705417\|20 | ···················· | ···················· | ···················· | ···················· | ···················· | ···················· | |
| W | 2168 hCoV-19/Wales/PHWC-491C57/2020\|EPI_ISL_705419\|2020 | ···················· | ···················· | ···················· | ···················· | ···················· | ···················· | |
| W | 2170 hCoV-19/Wales/PHWC-491AC6/2020\|EPI_ISL_706738\|2020 | ···················· | ···················· | ···················· | ···················· | ···················· | ···················· | |
| W | 2171 hCoV-19/Wales/PHWC-491AD5/2020\|EPI_ISL_706739\|2020 | ···················· | ···················· | ···················· | ···················· | ······T············· | ···················· | |
| W | 2172 hCoV-19/Wales/PHWC-491AE4/2020\|EPI_ISL_706740\|2020 | ···················· | ···················· | ···················· | ···················· | NNN················· | ···················· | 1 |
| W | 2175 hCoV-19/Wales/PHWC-491B1E/2020\|EPI_ISL_706743\|2020 | ···················· | ···················· | ···················· | ···················· | ···················· | ···················· | |
| W | 2176 hCoV-19/Wales/PHWC-491B2D/2020\|EPI_ISL_706744\|2020 | ···················· | ···················· | ···················· | ···················· | ···················· | ···················· | |
| W | 2177 hCoV-19/Wales/PHWC-491B3C/2020\|EPI_ISL_706745\|2020 | ···················· | ···················· | ···················· | ···················· | ···················· | ···················· | |
| W | 2181 hCoV-19/Wales/PHWC-491B78/2020\|EPI_ISL_706749\|2020 | ···················· | ···················· | ···················· | ···················· | ···················· | ···················· | |
| W | 2182 hCoV-19/USA/MN-MDH-2162/2020\|EPI_ISL_708500\|2020-1 | ···················· | ·A·················· | ···················· | ···················· | ···················· | ···················· | |
| W | 2183 hCoV-19/USA/MN-MDH-2163/2020\|EPI_ISL_708501\|2020-1 | ···················· | ·A·················· | ···················· | ···················· | ···················· | ···················· | |
| W | 2184 hCoV-19/USA/MN-MDH-2164/2020\|EPI_ISL_708502\|2020-1 | ···················· | ···················· | ···················· | ···················· | ···················· | ···················· | |
| W | 2185 hCoV-19/USA/MN-MDH-2165/2020\|EPI_ISL_708503\|2020-1 | ···················· | ···················· | ···················· | ···················· | ···················· | ···················· | |
| W | 2186 hCoV-19/USA/MN-MDH-2166/2020\|EPI_ISL_708504\|2020-0 | ···················· | ···················· | ···················· | ···················· | ···················· | ···················· | |
| W | 2187 hCoV-19/USA/CA-LACPHL-AD00005/2020\|EPI_ISL_708505\| | ···················· | ···················· | ···················· | ···················· | ···················· | ···················· | |
| W | 2188 hCoV-19/USA/CA-LACPHL-AD00010/2020\|EPI_ISL_708507\| | ···················· | ···················· | ···················· | ···················· | ···················· | ···················· | |
| W | 2189 hCoV-19/Ireland/MH-NVRL-20G47269/2020\|EPI_ISL_7085 | ···················· | ···················· | ···················· | ···················· | ···················· | ···················· | |
| W | 2190 hCoV-19/Ireland/D-NVRL-20G48650/2020\|EPI_ISL_70852 | ···················· | ···················A | ···················· | ···················· | ···················· | ···················· | 1 |
| W | 2191 hCoV-19/Brazil/SP-844R1/2020\|EPI_ISL_708529\|2020-0 | ···················· | ···················· | ···················· | ···················· | ···················· | ···················· | |
| W | 2192 hCoV-19/Brazil/SP-844R2/2020\|EPI_ISL_708530\|2020-1 | ···················· | ···················· | ···················· | ···················· | ···················· | ···················· | |
| W | 2193 hCoV-19/USA/MI-MDHHS-SC22544/2020\|EPI_ISL_708563\|2 | ···················· | ···················· | ···················· | ···················· | ···················· | ···················· | |
| W | 2194 hCoV-19/USA/MI-MDHHS-SC22466/2020\|EPI_ISL_708568\|2 | ···················· | ·············A······ | ···················· | ···················· | ···················· | ···················· | |
| W | 2195 hCoV-19/USA/MI-MDHHS-SC22467/2020\|EPI_ISL_708569\|2 | ···················· | ·············A······ | ···················· | ···················· | ···················· | ···················· | |
| W | 2196 hCoV-19/USA/MI-MDHHS-SC22468/2020\|EPI_ISL_708570\|2 | ···················· | ·············A······ | ···················· | ···················· | ···················· | ···················· | |
| W | 2197 hCoV-19/USA/MI-MDHHS-SC22469/2020\|EPI_ISL_708571\|2 | ···················· | ···················· | ···················· | ···················· | ···················· | ···················· | |
| W | 2198 hCoV-19/USA/MI-MDHHS-SC22470/2020\|EPI_ISL_708572\|2 | ···················· | ···················· | ···················· | ···················· | ···················· | ···················· | |
| W | 2199 hCoV-19/USA/MI-MDHHS-SC22472/2020\|EPI_ISL_708573\|2 | ···················· | ···················· | ···················· | ···················· | ···················· | ···················· | |
| W | 2200 hCoV-19/USA/MI-MDHHS-SC22473/2020\|EPI_ISL_708574\|2 | ···················· | ···················· | ···················· | ···················· | ···················· | ···················· | |
| W | 2201 hCoV-19/USA/MI-MDHHS-SC22474/2020\|EPI_ISL_708575\|2 | ···················· | ···················· | ···················· | ···················· | ···················· | ···················· | |
| W | 2202 hCoV-19/USA/MI-MDHHS-SC22475/2020\|EPI_ISL_708576\|2 | ···················· | ·············A······ | ···················· | ···················· | ···················· | ···················· | |
| W | 2203 hCoV-19/USA/MI-MDHHS-SC22476/2020\|EPI_ISL_708577\|2 | ···················· | ···················· | ···················· | ···················· | ···················· | ···················· | |
| W | 2204 hCoV-19/USA/MI-MDHHS-SC22477/2020\|EPI_ISL_708578\|2 | ···················· | ···················· | ···················· | ···················· | ···················· | ···················· | |
| W | 2205 hCoV-19/USA/MI-MDHHS-SC22478/2020\|EPI_ISL_708579\|2 | ···················· | ···················· | ···················· | ···················· | ···················· | ···················· | |
| W | 2206 hCoV-19/USA/MI-MDHHS-SC22479/2020\|EPI_ISL_708580\|2 | ···················· | ···················· | ···················· | ···················· | ···················· | ···················· | |
| W | 2207 hCoV-19/USA/MI-MDHHS-SC22480/2020\|EPI_ISL_708581\|2 | ···················· | ·············A······ | ···················· | ···················· | ···················· | ···················· | |
| W | 2208 hCoV-19/USA/MI-MDHHS-SC22481/2020\|EPI_ISL_708582\|2 | ···················· | ···················· | ···················· | ···················· | ···················· | ···················· | |
| W | 2209 hCoV-19/USA/MI-MDHHS-SC22482/2020\|EPI_ISL_708583\|2 | ···················· | ···················· | ···················· | ···················· | ···················· | ···················· | |
| W | 2210 hCoV-19/USA/MI-MDHHS-SC22483/2020\|EPI_ISL_708584\|2 | ···················· | ·············A······ | ···················· | ···················· | ···················· | ···················· | |
| W | 2211 hCoV-19/USA/MI-MDHHS-SC22484/2020\|EPI_ISL_708585\|2 | ···················· | ···················· | ···················· | ···················· | ···················· | ···················· | |
| W | 2212 hCoV-19/USA/MI-MDHHS-SC22485/2020\|EPI_ISL_708586\|2 | ···················· | ···················· | ···················· | ···················· | ···················· | ···················· | |
| W | 2213 hCoV-19/USA/MI-MDHHS-SC22486/2020\|EPI_ISL_708587\|2 | ···················· | ···················· | ···················· | ···················· | ···················· | ···················· | |
| W | 2214 hCoV-19/USA/MI-MDHHS-SC22487/2020\|EPI_ISL_708588\|2 | ···················· | ···················· | ···················· | ···················· | ···················· | ···················· | |
| W | 2215 hCoV-19/USA/MI-MDHHS-SC22488/2020\|EPI_ISL_708589\|2 | ···················· | ···················· | ···················· | ···················· | ···················· | ···················· | |
| W | 2216 hCoV-19/USA/MI-MDHHS-SC22489/2020\|EPI_ISL_708590\|2 | ···················· | ···················· | ···················· | ···················· | ···················· | ···················· | |
| W | 2217 hCoV-19/USA/MI-MDHHS-SC22490/2020\|EPI_ISL_708591\|2 | ···················· | ·············A······ | ···················· | ···················· | ···················· | ···················· | |
| W | 2218 hCoV-19/USA/MI-MDHHS-SC22491/2020\|EPI_ISL_708592\|2 | ···················· | ·············A······ | ···················· | ···················· | ···················· | ···················· | |
| W | 2219 hCoV-19/USA/MI-MDHHS-SC22492/2020\|EPI_ISL_708593\|2 | ···················· | ···················· | ···················· | ···················· | ···················· | ···················· | |
| W | 2220 hCoV-19/USA/MI-MDHHS-SC22493/2020\|EPI_ISL_708594\|2 | ···················· | ···················· | ···················· | ···················· | ···················· | ···················· | |
| W | 2221 hCoV-19/USA/MI-MDHHS-SC22494/2020\|EPI_ISL_708595\|2 | ···················· | ···················· | ···················· | ···················· | ···················· | ···················· | |
| W | 2222 hCoV-19/USA/MI-MDHHS-SC22495/2020\|EPI_ISL_708596\|2 | ···················· | ···················· | ···················· | ···················· | ···················· | ···················· | |
| W | 2223 hCoV-19/USA/MI-MDHHS-SC22496/2020\|EPI_ISL_708597\|2 | ···················· | ···················· | ···················· | ···················· | ···················· | ···················· | |
| W | 2224 hCoV-19/USA/MI-MDHHS-SC22539/2020\|EPI_ISL_708598\|2 | ···················· | ···················· | ···················· | ···················· | ···················· | ···················· | |
| W | 2225 hCoV-19/USA/MI-MDHHS-SC22540/2020\|EPI_ISL_708599\|2 | ···················· | ·············A······ | ···················· | ···················· | ···················· | ···················· | |
| W | 2226 hCoV-19/England/QEUH-BE40DA/2020\|EPI_ISL_709800\|20 | ···················· | ···················· | ···················· | ···················· | ···················· | ···················· | |
| W | 2227 hCoV-19/England/MILK-BE7365/2020\|EPI_ISL_709802\|20 | ···················· | ···················· | ···················· | ···················· | ······T············· | ···················· | |
| W | 2228 hCoV-19/England/CAMC-BDD001/2020\|EPI_ISL_709805\|20 | ···················· | ···················· | ···················· | ···················· | ···················· | ···················· | |
| W | 2229 hCoV-19/England/QEUH-BE5798/2020\|EPI_ISL_709806\|20 | ···················· | ···················· | ···················· | ···················· | ···················· | ···················· | |
| W | 2230 hCoV-19/England/CAMC-BDCFE7/2020\|EPI_ISL_709807\|20 | ···················· | ···················· | ···················· | ···················· | ···················· | ···················· | |
| W | 2231 hCoV-19/England/CAMC-BDD03E/2020\|EPI_ISL_709808\|20 | ···················· | ·A·················· | ···················· | ···················· | ···················· | ···A················ | |
| W | 2232 hCoV-19/England/QEUH-BE40AD/2020\|EPI_ISL_709809\|20 | ···················· | ···················· | ···················· | ···················· | ···················· | ···················· | |
| W | 2233 hCoV-19/England/CAMC-BDD07A/2020\|EPI_ISL_709810\|20 | ···················· | ···················· | ···················· | ···················· | ···················· | ···················· | |
| W | 2234 hCoV-19/England/QEUH-BE4131/2020\|EPI_ISL_709811\|20 | ···················· | ···················· | ···················· | ···················· | ···················· | ···················· | |
| W | 2235 hCoV-19/England/QEUH-BE57B6/2020\|EPI_ISL_709812\|20 | ···················· | ···················· | ···················· | ···················· | ···················· | ···················· | |
| W | 2236 hCoV-19/England/CAMC-BDD04D/2020\|EPI_ISL_709813\|20 | ···················· | ···················· | ···················· | ···················· | ···················· | ···················· | |
| W | 2237 hCoV-19/England/CAMC-BDD010/2020\|EPI_ISL_709814\|20 | ···················· | ···················· | ···················· | ···················· | ···················· | ···················· | |
| W | 2238 hCoV-19/England/MILK-BE715C/2020\|EPI_ISL_709815\|20 | ···················· | ···················· | ···················· | ···················· | ···················· | ···················· | |
| W | 2239 hCoV-19/England/QEUH-BE4070/2020\|EPI_ISL_709816\|20 | ···················· | ···················· | ···················· | ···················· | ···················· | ···················· | |
| W | 2240 hCoV-19/England/CAMC-BE036C/2020\|EPI_ISL_709817\|20 | ···················· | ···················· | ···················· | ···················· | ···················· | ···················· | |
| W | 2241 hCoV-19/England/QEUH-BE40BC/2020\|EPI_ISL_709818\|20 | ···················· | ···················· | ···················· | ···················· | ···················· | ···················· | |
| W | 2242 hCoV-19/England/MILK-BE71D4/2020\|EPI_ISL_709819\|20 | ···················· | ···················· | ···················· | ···················· | ······T············· | ···················· | |
| W | 2243 hCoV-19/England/QEUH-BE4140/2020\|EPI_ISL_709820\|20 | ···················· | ···················· | ···················· | ···················· | ···················· | ···················· | |
| W | 2244 hCoV-19/England/CAMC-BE033F/2020\|EPI_ISL_709822\|20 | ···················· | ···················· | ···················· | ···················· | ···················· | ···················· | |
| W | 2245 hCoV-19/England/MILK-BE7040/2020\|EPI_ISL_709823\|20 | ···················· | ·A·················· | ···················· | ···················· | ···················· | ···A················ | |
| W | 2246 hCoV-19/England/MILK-BE7031/2020\|EPI_ISL_709824\|20 | ···················· | ···················· | ···················· | ···················· | ···················· | ···················· | |
| W | 2247 hCoV-19/England/QEUH-BE57C5/2020\|EPI_ISL_709826\|20 | ···················· | ···················· | ···················· | ···················· | ···················· | ···················· | |
| W | 2248 hCoV-19/England/CAMC-BDCC86/2020\|EPI_ISL_709827\|20 | ···················· | ···················· | ···················· | ···················· | ···················· | ···················· | |

```
W    2249 hCoV-19/England/MILK-BE71A7/2020|EPI_ISL_709828|20   ····················  ····················  ·····················  ····················  ····················  ····················
W    2250 hCoV-19/England/MILK-BE7013/2020|EPI_ISL_709830|20   ····················  ····················  ·····················  ····················  ····················  ····················
W    2251 hCoV-19/England/QEUH-BE409E/2020|EPI_ISL_709831|20   ····················  ····················  ·····················  ····················  ····················  ····················
W    2252 hCoV-19/England/QEUH-BE580E/2020|EPI_ISL_709832|20   ····················  ····················  ·····················  ····················  ····················  T···················
W    2253 hCoV-19/England/MILK-BE7374/2020|EPI_ISL_709833|20   ····················  ····················  ·····················  ····················  ·····T··············  ····················
W    2254 hCoV-19/England/QEUH-BE4061/2020|EPI_ISL_709834|20   ····················  ····················  ·····················  ····················  ····················  ····················
W    2255 hCoV-19/England/QEUH-BE4113/2020|EPI_ISL_709835|20   ····················  ····················  ·····················  ····················  ····················  ····················
W    2256 hCoV-19/England/CAMC-BDCC77/2020|EPI_ISL_709836|20   ····················  ····················  ·····················  ····················  ····················  ····················
W    2257 hCoV-19/England/MILK-BE6FCC/2020|EPI_ISL_709838|20   ····················  ····················  ·····················  ····················  ····················  ····················
W    2258 hCoV-19/England/MILK-BE71B6/2020|EPI_ISL_709839|20   ····················  ····················  ·····················  ····················  ····················  ····················
W    2259 hCoV-19/England/QEUH-BE416E/2020|EPI_ISL_709840|20   ····················  ····················  ·····················  ····················  ····················  ····················
W    2260 hCoV-19/England/MILK-BE7198/2020|EPI_ISL_709841|20   ····················  ····················  ·····················  ····················  ····················  ····················
W    2261 hCoV-19/England/CAMC-BE037B/2020|EPI_ISL_709843|20   ····················  ····················  ·····················  ····················  ····················  ····················
W    2262 hCoV-19/England/QEUH-BE415F/2020|EPI_ISL_709845|20   ····················  ····················  ·····················  ····················  ····················  ····················
W    2263 hCoV-19/England/QEUH-BE4043/2020|EPI_ISL_709848|20   ····················  ····················  ·····················  ····················  ····················  ····················
W    2264 hCoV-19/England/QEUH-BE40CB/2020|EPI_ISL_709849|20   ····················  ····················  ·····················  ····················  ····················  ····················
W    2265 hCoV-19/England/CAMC-BE0311/2020|EPI_ISL_709850|20   ····················  ····················  ·····················  ····················  ····················  ····················
W    2266 hCoV-19/England/QEUH-BE577A/2020|EPI_ISL_709851|20   ····················  ····················  ·····················  ····················  ····················  ····················
W    2267 hCoV-19/England/MILK-BE6FEA/2020|EPI_ISL_709852|20   ····················  ····················  ·····················  ····················  ····················  ····················
W    2268 hCoV-19/England/MILK-BE71C5/2020|EPI_ISL_709853|20   ····················  ····················  ·····················  ····················  ····················  ····················
W    2269 hCoV-19/England/CAMC-BDCC59/2020|EPI_ISL_709854|20   ····················  ····················  ·····················  ····················  ·····T··············  ····················
W    2270 hCoV-19/England/QEUH-BE4122/2020|EPI_ISL_709855|20   ····················  ····················  ·····················  ····················  ····················  ····················
W    2271 hCoV-19/England/MILK-BE7347/2020|EPI_ISL_709856|20   ····················  ····················  ·····················  ····················  ····················  ····················
W    2272 hCoV-19/England/QEUH-BE40F8/2020|EPI_ISL_709857|20   ····················  ····················  ·····················  ····················  ·····T··············  ····················
W    2273 hCoV-19/England/QEUH-BE408F/2020|EPI_ISL_709858|20   ····················  ····················  ·····················  ····················  ····················  ····················
W    2274 hCoV-19/England/QEUH-BE57D4/2020|EPI_ISL_709859|20   ····················  ····················  ·····················  ····················  ····················  ····················
W    2275 hCoV-19/England/MILK-BE717A/2020|EPI_ISL_709860|20   ····················  ·A··················  ·····················  ····················  ····················  ··A·················
W    2276 hCoV-19/England/QEUH-BE4052/2020|EPI_ISL_709861|20   ····················  ····················  ·····················  ····················  ····················  ····················
W    2277 hCoV-19/England/MILK-BE6FDB/2020|EPI_ISL_709863|20   ····················  ····················  ·····················  ····················  ····················  ····················
W    2278 hCoV-19/England/CAMC-BDCFC9/2020|EPI_ISL_709864|20   ····················  ····················  ·····················  ····················  ·····T··············  ····················
W    2279 hCoV-19/England/MILK-BE6FF9/2020|EPI_ISL_709866|20   ····················  ····················  ·····················  ····················  ····················  ····················
W    2280 hCoV-19/England/CAMC-BDCFBA/2020|EPI_ISL_709867|20   ····················  ····················  ·····················  ····················  ····················  ····················
W    2281 hCoV-19/England/MILK-BE7022/2020|EPI_ISL_709868|20   ····················  ·A··················  ·····················  ····················  ····················  ····················
W    2282 hCoV-19/England/CAMC-BDD089/2020|EPI_ISL_709869|20   ····················  ····················  ·····················  ····················  ····················  ····················
W    2283 hCoV-19/England/CAMC-BE0302/2020|EPI_ISL_709870|20   ····················  ····················  ·····················  ····················  ····················  ····················
W    2284 hCoV-19/England/MILK-BE7189/2020|EPI_ISL_709871|20   ····················  ····················  ·····················  ····················  ····················  ····················
W    2285 hCoV-19/England/MILK-BE71E3/2020|EPI_ISL_709872|20   ····················  ····················  ·····················  ····················  ····················  ····················
W    2286 hCoV-19/England/QEUH-BE57F2/2020|EPI_ISL_709875|20   ····················  ····················  ·····················  ····················  ····················  ····················
W    2287 hCoV-19/England/CAMC-BDCFF6/2020|EPI_ISL_709876|20   ····················  ····················  ·····················  ····················  ····················  ····················
W    2288 hCoV-19/England/CAMC-BE035D/2020|EPI_ISL_709877|20   ····················  ····················  ·····················  ····················  ····················  ····················
W    2289 hCoV-19/England/MILK-BE6FBD/2020|EPI_ISL_709879|20   ····················  ····················  ·····················  ····················  ····················  ····················
W    2290 hCoV-19/England/CAMC-BE02E7/2020|EPI_ISL_709880|20   ····················  ····················  ·····················  ····················  ····················  ····················
W    2291 hCoV-19/England/MILK-BE7356/2020|EPI_ISL_709881|20   ····················  ····················  ·····················  ····················  ····················  ····················
W    2292 hCoV-19/England/MILK-BAC469/2020|EPI_ISL_709882|20   ····················  ····················  ·····················  ····················  ·····T··············  ····················
W    2293 hCoV-19/England/MILK-B94EA2/2020|EPI_ISL_709883|20   ····················  ····················  ·····················  ····················  ····················  ····················
W    2294 hCoV-19/England/MILK-B94E93/2020|EPI_ISL_709884|20   ····················  ····················  ·····················  ····················  ····················  ····················
W    2295 hCoV-19/England/MILK-B8820F/2020|EPI_ISL_709885|20   ····················  ····················  ·····················  ····················  ····················  ····················
W    2296 hCoV-19/England/MILK-B8C03E/2020|EPI_ISL_709886|20   ····················  ····················  ·····················  ····················  ·····T··············  ····················
W    2297 hCoV-19/England/MILK-B881F3/2020|EPI_ISL_709887|20   ····················  ····················  ·····················  ····················  ····················  ····················
W    2298 hCoV-19/England/MILK-B8821E/2020|EPI_ISL_709888|20   ····················  ····················  ·····················  ····················  ····················  ····················
W    2299 hCoV-19/England/MILK-B89F25/2020|EPI_ISL_709890|20   ····················  ····················  ·····················  ····················  ····················  ····················
W    2300 hCoV-19/England/MILK-B88E74/2020|EPI_ISL_709891|20   ····················  ····················  ·····················  ····················  ····················  ····················
W    2301 hCoV-19/England/MILK-B8C906/2020|EPI_ISL_709893|20   ····················  ····················  ·····················  ····················  ····················  ····················
W    2302 hCoV-19/England/MILK-B94B14/2020|EPI_ISL_709895|20   ····················  ····················  ·····················  ····················  ····················  ····················
W    2303 hCoV-19/England/MILK-B967F0/2020|EPI_ISL_709896|20   ····················  ····················  ·····················  ····················  ····················  ····················
W    2304 hCoV-19/Northern Ireland/MILK-B947A7/2020|EPI_ISL_   ····················  ····················  ·····················  ····················  ····················  ····················
W    2305 hCoV-19/USA/AZ-TG276579/2020|EPI_ISL_694113|2020-0   ····················  ····················  ·····················  ····················  ····················  ····················
W    2306 hCoV-19/USA/AZ-TG276586/2020|EPI_ISL_694115|2020-0   ····················  ····················  ·····················  ····················  ····················  ····················
W    2307 hCoV-19/USA/AZ-TG276592/2020|EPI_ISL_694117|2020-0   ····················  ····················  ·····················  ····················  ····················  ····················
W    2308 hCoV-19/USA/AZ-TG276740/2020|EPI_ISL_694120|2020-0   ····················  ····················  ·····················  ····················  ····················  ····················
W    2309 hCoV-19/USA/AZ-TG276752/2020|EPI_ISL_694124|2020-0   ····················  ····················  ·····················  ····················  ····················  ····················
W    2310 hCoV-19/USA/AZ-TG276754/2020|EPI_ISL_694125|2020-0   ····················  ····················  ·····················  ····················  ····················  ····················
W    2311 hCoV-19/USA/AZ-TG276756/2020|EPI_ISL_694126|2020-0   ····················  ····················  ·····················  ····················  ····················  ····················
W    2312 hCoV-19/USA/AZ-TG276758/2020|EPI_ISL_694127|2020-0   ····················  ····················  ·····················  ····················  ····················  ····················
W    2313 hCoV-19/USA/AZ-TG276762/2020|EPI_ISL_694128|2020-0   ····················  ····················  ·····················  ····················  ····················  ····················
W    2314 hCoV-19/USA/AZ-TG276772/2020|EPI_ISL_694132|2020-0   ····················  ····················  ·····················  ····················  ····················  ····················
W    2315 hCoV-19/USA/AZ-TG276776/2020|EPI_ISL_694134|2020-0   ····················  ····················  ·····················  ····················  ····················  ····················
W    2316 hCoV-19/USA/AZ-TG277985/2020|EPI_ISL_694139|2020-0   ····················  ····················  ·····················  ····················  ····················  ····················
W    2317 hCoV-19/USA/AZ-TG278043/2020|EPI_ISL_694140|2020-0   ····················  ····················  ·····················  ····················  ····················  ····················
W    2318 hCoV-19/USA/AZ-TG278298/2020|EPI_ISL_694141|2020-0   ····················  ····················  ·····················  ····················  ····················  ····················
W    2319 hCoV-19/USA/AZ-TG279396/2020|EPI_ISL_694143|2020-0   ····················  ····················  ·····················  ····················  ····················  ····················
W    2320 hCoV-19/USA/AZ-TG279676/2020|EPI_ISL_694144|2020-0   ····················  ····················  ·····················  ····················  ····················  ····················
W    2321 hCoV-19/USA/AZ-TG280089/2020|EPI_ISL_694145|2020-0   ····················  ····················  ·····················  ····················  ····················  ····················
W    2322 hCoV-19/USA/AZ-TG280188/2020|EPI_ISL_694146|2020-0   ····················  ····················  ·····················  ····················  ····················  ····················
W    2323 hCoV-19/USA/AZ-TG280790/2020|EPI_ISL_694149|2020-0   ····················  ····················  ·····················  ····················  ····················  ····················
W    2324 hCoV-19/USA/AZ-TG280794/2020|EPI_ISL_694151|2020-0   ····················  ····················  ·····················  ····················  ····················  ····················
W    2325 hCoV-19/USA/AZ-TG280795/2020|EPI_ISL_694152|2020-0   ····················  ····················  ·····················  ····················  ····················  ····················
W    2326 hCoV-19/USA/AZ-TG280801/2020|EPI_ISL_694155|2020-0   ····················  ····················  ·····················  ····················  ····················  ····················
W    2327 hCoV-19/USA/AZ-TG280802/2020|EPI_ISL_694156|2020-0   ····················  ····················  ·····················  ····················  ····················  ····················
W    2328 hCoV-19/USA/AZ-TG280803/2020|EPI_ISL_694157|2020-0   ····················  ····················  ·····················  ····················  ····················  ····················
```

| | | | | | | | |
|---|---|---|---|---|---|---|---|
| W | 2329 hCoV-19/USA/AZ-TG280806/2020\|EPI_ISL_694159\|2020-0 | ···················· | ···················· | ···················· | ···················· | ···················· | ···················· |
| W | 2330 hCoV-19/USA/AZ-TG280811/2020\|EPI_ISL_694162\|2020-0 | ···················· | ···················· | ···················· | ···················· | ···················· | ···················· |
| W | 2331 hCoV-19/USA/AZ-TG547248/2020\|EPI_ISL_695446\|2020-0 | ···················· | ···················· | ···················· | ···················· | ···················· | ···················· |
| W | 2332 hCoV-19/USA/AZ-TG547255/2020\|EPI_ISL_695451\|2020-0 | ···················· | ···················· | ···················· | ···················· | ···················· | ···················· |
| W | 2333 hCoV-19/USA/AZ-TG547256/2020\|EPI_ISL_695452\|2020-0 | ···················· | ···················· | ···················· | ···················· | ···················· | ···················· |
| W | 2334 hCoV-19/USA/AZ-TG547260/2020\|EPI_ISL_695454\|2020-0 | ···················· | ···················· | ···················· | ···················· | ···················· | ···················· |
| W | 2335 hCoV-19/USA/AZ-TG547261/2020\|EPI_ISL_695455\|2020-0 | ···················· | ···················· | ···················· | ···················· | ···················· | ···················· |
| W | 2336 hCoV-19/USA/AZ-TG547263/2020\|EPI_ISL_695456\|2020-0 | ···················· | ···················· | ···················· | ···················· | ···················· | ···················· |
| W | 2337 hCoV-19/USA/AZ-TG547274/2020\|EPI_ISL_695458\|2020-0 | ···················· | ···················· | ···················· | ···················· | ···················· | ···················· |
| W | 2338 hCoV-19/USA/AZ-TG547275/2020\|EPI_ISL_695459\|2020-0 | ···················· | ···················· | ···················· | ···················· | ···················· | ···················· |
| W | 2339 hCoV-19/USA/AZ-TG547283/2020\|EPI_ISL_695462\|2020-0 | ···················· | ·A·················· | ···················· | ···················· | ···················· | ···················· |
| W | 2340 hCoV-19/USA/AZ-TG547288/2020\|EPI_ISL_695463\|2020-0 | ···················· | ···················· | ···················· | ···················· | ···················· | ···················· |
| W | 2341 hCoV-19/USA/AZ-TG547290/2020\|EPI_ISL_695465\|2020-0 | ···················· | ·A·················· | ···················· | ···················· | ···················· | ···················· |
| W | 2342 hCoV-19/USA/AZ-TG547291/2020\|EPI_ISL_695466\|2020-0 | ···················· | ·A·················· | ···················· | ···················· | ···················· | ···················· |
| W | 2343 hCoV-19/USA/AZ-TG547296/2020\|EPI_ISL_695470\|2020-0 | ···················· | ·A·················· | ···················· | ···················· | ···················· | ···················· |
| W | 2344 hCoV-19/USA/AZ-TG547300/2020\|EPI_ISL_695472\|2020-0 | ···················· | ·A·················· | ···················· | ···················· | ···················· | ···················· |
| W | 2345 hCoV-19/USA/AZ-TG547301/2020\|EPI_ISL_695473\|2020-0 | ···················· | ·A·················· | ···················· | ···················· | ···················· | ···················· |
| W | 2346 hCoV-19/USA/AZ-TG547306/2020\|EPI_ISL_695476\|2020-0 | ···················· | ···················· | ···················· | ···················· | ···················· | ···················· |
| W | 2347 hCoV-19/USA/AZ-TG547311/2020\|EPI_ISL_695478\|2020-0 | ···················· | ···················· | ···················· | ···················· | ···················· | ···················· |
| W | 2348 hCoV-19/USA/AZ-TG547316/2020\|EPI_ISL_695479\|2020-0 | ···················· | ···················· | ···················· | ···················· | ···················· | ···················· |
| W | 2349 hCoV-19/USA/AZ-TG547322/2020\|EPI_ISL_695480\|2020-0 | ···················· | ·A·················· | ···················· | ···················· | ···················· | ···················· |
| W | 2350 hCoV-19/USA/AZ-TG547325/2020\|EPI_ISL_695481\|2020-0 | ···················· | ···················· | ···················· | ···················· | ···················· | ···················· |
| W | 2351 hCoV-19/USA/AZ-TG547329/2020\|EPI_ISL_695482\|2020-0 | ···················· | ·A·················· | ···················· | ···················· | ···················· | ···················· |
| W | 2352 hCoV-19/USA/AZ-TG547333/2020\|EPI_ISL_695484\|2020-0 | ···················· | ·A·················· | ···················· | ···················· | ···················· | ···················· |
| W | 2353 hCoV-19/USA/AZ-TG547337/2020\|EPI_ISL_695486\|2020-0 | ···················· | ·A·················· | ···················· | ···················· | ···················· | ···················· |
| W | 2354 hCoV-19/USA/AZ-TG547339/2020\|EPI_ISL_695488\|2020-0 | ···················· | ·A·················· | ···················· | ···················· | ···················· | ···················· |
| W | 2355 hCoV-19/USA/AZ-TG547340/2020\|EPI_ISL_695489\|2020-0 | ···················· | ·A·················· | ···················· | ···················· | ···················· | ···················· |
| W | 2356 hCoV-19/USA/AZ-TG547348/2020\|EPI_ISL_695496\|2020-0 | ···················· | ···················· | ···················· | ···················· | ···················· | ···················· |
| W | 2357 hCoV-19/USA/AZ-TG547352/2020\|EPI_ISL_695498\|2020-0 | ···················· | ·A·················· | ···················· | ···················· | ···················· | ···················· |
| W | 2358 hCoV-19/United Arab Emirates/0861/2020\|EPI_ISL_699 | ···················· | ···················· | ···················· | ···················· | ···················· | ···················· |
| W | 2359 hCoV-19/United Arab Emirates/0862/2020\|EPI_ISL_699 | ···················· | ···················· | ···················· | ···················· | ···················· | ···················· |
| W | 2360 hCoV-19/United Arab Emirates/0863/2020\|EPI_ISL_699 | ···················· | ···················· | ···················· | ···················· | ···················· | ···················· |
| W | 2361 hCoV-19/United Arab Emirates/0864/2020\|EPI_ISL_699 | ···················· | ···················· | ···················· | ···················· | ···················· | ···················· |
| W | 2362 hCoV-19/United Arab Emirates/0865/2020\|EPI_ISL_699 | ···················· | ···················· | ···················· | ···················· | ···················· | ···················· |
| W | 2363 hCoV-19/United Arab Emirates/0866/2020\|EPI_ISL_699 | ···················· | ···················· | ···················· | ···················· | ···················· | ···················· |
| W | 2364 hCoV-19/United Arab Emirates/0867/2020\|EPI_ISL_699 | ···················· | ···················· | ···················· | ···················· | ···················· | ···················· |
| W | 2365 hCoV-19/United Arab Emirates/0868/2020\|EPI_ISL_699 | ···················· | ···················· | ···················· | ···················· | ···················· | ···················· |
| W | 2366 hCoV-19/United Arab Emirates/0882/2020\|EPI_ISL_699 | ···················· | ···················· | ···················· | ···················· | ···················· | ···················· |
| W | 2367 hCoV-19/United Arab Emirates/0892/2020\|EPI_ISL_699 | ···················· | ···················· | ···················· | ···················· | ···················· | ···················· |
| W | 2368 hCoV-19/United Arab Emirates/0893/2020\|EPI_ISL_699 | ···················· | ···················· | ···················· | ···················· | ···················· | ···················· |
| W | 2369 hCoV-19/United Arab Emirates/0894/2020\|EPI_ISL_699 | ···················· | ···················· | ···················· | ···················· | ···················· | ···················· |
| W | 2370 hCoV-19/United Arab Emirates/0895/2020\|EPI_ISL_699 | ···················· | ···················· | ···················· | ···················· | ···················· | ···················· |
| W | 2371 hCoV-19/United Arab Emirates/0896/2020\|EPI_ISL_699 | ···················· | ···················· | ···················· | ···················· | ···················· | ···················· |
| W | 2372 hCoV-19/United Arab Emirates/0897/2020\|EPI_ISL_699 | ···················· | ···················· | ···················· | ···················· | ···················· | ···················· |
| W | 2373 hCoV-19/United Arab Emirates/0898/2020\|EPI_ISL_699 | ···················· | ···················· | ···················· | ···················· | ···················· | ···················· |
| W | 2374 hCoV-19/United Arab Emirates/0899/2020\|EPI_ISL_699 | ···················· | ···················· | ···················· | ···················· | ···················· | ···················· |
| W | 2375 hCoV-19/United Arab Emirates/0900/2020\|EPI_ISL_699 | ···················· | ···················· | ···················· | ···················· | ···················· | ···················· |
| W | 2376 hCoV-19/United Arab Emirates/0901/2020\|EPI_ISL_699 | ···················· | ···················· | ···················· | ···················· | ···················· | ···················· |
| W | 2377 hCoV-19/United Arab Emirates/0902/2020\|EPI_ISL_699 | ···················· | ···················· | ···················· | ···················· | ···················· | ···················· |
| W | 2378 hCoV-19/United Arab Emirates/0903/2020\|EPI_ISL_699 | ···················· | ···················· | ···················· | ···················· | ···················· | ···················· |
| W | 2379 hCoV-19/United Arab Emirates/0904/2020\|EPI_ISL_699 | ···················· | ···················· | ···················· | ···················· | ···················· | ···················· |
| W | 2380 hCoV-19/United Arab Emirates/0905/2020\|EPI_ISL_699 | ···················· | ···················· | ···················· | ···················· | ···················· | ···················· |
| W | 2381 hCoV-19/United Arab Emirates/0906/2020\|EPI_ISL_699 | ···················· | ·A·················· | ···················· | ···················· | ···················· | ··A················· |
| W | 2382 hCoV-19/United Arab Emirates/0907/2020\|EPI_ISL_699 | ···················· | ···················· | ···················· | ···················· | ···················· | ···················· |
| W | 2383 hCoV-19/United Arab Emirates/0908/2020\|EPI_ISL_699 | ···················· | ···················· | ···················· | ···················· | ···················· | ···················· |
| W | 2384 hCoV-19/United Arab Emirates/0909/2020\|EPI_ISL_699 | ···················· | ···················· | ···················· | ···················· | ···················· | ···················· |
| W | 2385 hCoV-19/United Arab Emirates/0910/2020\|EPI_ISL_699 | ···················· | ···················· | ···················· | ···················· | ···················· | ···················· |
| W | 2386 hCoV-19/United Arab Emirates/0911/2020\|EPI_ISL_699 | ···················· | ···················· | ···················· | ···················· | ···················· | ···················· |
| W | 2387 hCoV-19/United Arab Emirates/0912/2020\|EPI_ISL_699 | ···················· | ···················· | ···················· | ···················· | ···················· | ···················· |
| W | 2388 hCoV-19/United Arab Emirates/0913/2020\|EPI_ISL_699 | ···················· | ···················· | ···················· | ···················· | ···················· | ···················· |
| W | 2389 hCoV-19/United Arab Emirates/0914/2020\|EPI_ISL_699 | ···················· | ···················· | ···················· | ···················· | ···················· | ···················· |
| W | 2390 hCoV-19/United Arab Emirates/0915/2020\|EPI_ISL_699 | ···················· | ···················· | ···················· | ···················· | ···················· | ···················· |
| W | 2391 hCoV-19/United Arab Emirates/0916/2020\|EPI_ISL_699 | ···················· | ···················· | ···················· | ···················· | ···················· | ···················· |
| W | 2392 hCoV-19/United Arab Emirates/0917/2020\|EPI_ISL_699 | ···················· | ···················· | ···················· | ···················· | ···················· | ···············A···· |
| W | 2393 hCoV-19/United Arab Emirates/0918/2020\|EPI_ISL_699 | ···················· | ···················· | ···················· | ···················· | ···················· | ···············A···· |
| W | 2394 hCoV-19/United Arab Emirates/0919/2020\|EPI_ISL_699 | ···················· | ···················· | ···················· | ···················· | ···················· | ···················· |
| W | 2395 hCoV-19/United Arab Emirates/0920/2020\|EPI_ISL_699 | ···················· | ···················· | ···················· | ···················· | ···················· | ···················· |
| W | 2396 hCoV-19/United Arab Emirates/0921/2020\|EPI_ISL_699 | ···················· | ···················· | ···················· | ···················· | ···················· | ···················· |
| W | 2397 hCoV-19/United Arab Emirates/0922/2020\|EPI_ISL_699 | ···················· | ···················· | ···················· | ···················· | ···················· | ···················· |
| W | 2398 hCoV-19/United Arab Emirates/0923/2020\|EPI_ISL_699 | ···················· | ···················· | ···················· | ···················· | ···················· | ···················· |
| W | 2399 hCoV-19/United Arab Emirates/0924/2020\|EPI_ISL_699 | ···················· | ···················· | ···················· | ···················· | ···················· | ···················· |
| W | 2400 hCoV-19/United Arab Emirates/0925/2020\|EPI_ISL_699 | ···················· | ···················· | ···················· | ···················· | ···················· | ···················· |
| W | 2401 hCoV-19/United Arab Emirates/0926/2020\|EPI_ISL_699 | ···················· | ···················· | ···················· | ···················· | ···················· | ···················· |
| W | 2402 hCoV-19/United Arab Emirates/0927/2020\|EPI_ISL_699 | ···················· | ···················· | ···················· | ···················· | ···················· | ···················· |
| W | 2403 hCoV-19/United Arab Emirates/0928/2020\|EPI_ISL_699 | ···················· | ·A·················· | ···················· | ···················· | ···················· | ···················· |
| W | 2404 hCoV-19/United Arab Emirates/0929/2020\|EPI_ISL_699 | ···················· | ···················· | ···················· | ···················· | ···················· | ···················· |
| W | 2405 hCoV-19/United Arab Emirates/0930/2020\|EPI_ISL_699 | ···················· | ···················· | ···················· | ···················· | ···················· | ···················· |
| W | 2406 hCoV-19/United Arab Emirates/0932/2020\|EPI_ISL_699 | ···················· | ···················· | ···················· | ···················· | ···················· | ···················· |
| W | 2407 hCoV-19/United Arab Emirates/0934/2020\|EPI_ISL_699 | ···················· | ···················· | ···················· | ···················· | ···················· | ···················· |
| W | 2408 hCoV-19/United Arab Emirates/0935/2020\|EPI_ISL_699 | ···················· | ···················· | ···················· | ···················· | ···················· | ···················· |

| | | | | | | | | |
|---|---|---|---|---|---|---|---|---|
| W | 2409 | hCoV-19/United Arab Emirates/0936/2020\|EPI_ISL_699 | .................... | .................... | .................... | .................... | .................... | .................... |
| W | 2410 | hCoV-19/United Arab Emirates/0937/2020\|EPI_ISL_699 | .................... | .................... | .................... | .................... | .................... | .................... |
| W | 2411 | hCoV-19/United Arab Emirates/0938/2020\|EPI_ISL_699 | .................... | .................... | .................... | .................... | .................... | .................... |
| W | 2412 | hCoV-19/United Arab Emirates/0939/2020\|EPI_ISL_699 | .................... | .................... | .................... | .................... | .................... | .................... |
| W | 2413 | hCoV-19/United Arab Emirates/0940/2020\|EPI_ISL_699 | .................... | .................... | .................... | .................... | .................... | .................... |
| W | 2414 | hCoV-19/United Arab Emirates/0941/2020\|EPI_ISL_699 | .................... | .................... | .................... | .................... | .................... | .................... |
| W | 2415 | hCoV-19/United Arab Emirates/0942/2020\|EPI_ISL_699 | .................... | .................... | .................... | .................... | .................... | .................... |
| W | 2416 | hCoV-19/United Arab Emirates/0943/2020\|EPI_ISL_699 | .................... | .................... | .................... | .................... | .................... | .................... |
| W | 2417 | hCoV-19/United Arab Emirates/0944/2020\|EPI_ISL_699 | .................... | .................... | .................... | .................... | .................... | .................... |
| W | 2418 | hCoV-19/United Arab Emirates/0945/2020\|EPI_ISL_699 | .................... | .................... | .................... | .................... | .................... | .................... |
| W | 2419 | hCoV-19/United Arab Emirates/0946/2020\|EPI_ISL_699 | .................... | .................... | .................... | .................... | .................... | .................... |
| W | 2420 | hCoV-19/United Arab Emirates/0947/2020\|EPI_ISL_699 | .................... | .................... | .................... | .................... | .................... | .................... |
| W | 2421 | hCoV-19/United Arab Emirates/0948/2020\|EPI_ISL_699 | .................... | .................... | .................... | .................... | .................... | ...............A.... |
| W | 2422 | hCoV-19/United Arab Emirates/0949/2020\|EPI_ISL_699 | .................... | .................... | .................... | .................... | .................... | ...............A.... |
| W | 2423 | hCoV-19/United Arab Emirates/0950/2020\|EPI_ISL_699 | .................... | .................... | .................... | .................... | .................... | .................... |
| W | 2424 | hCoV-19/United Arab Emirates/0951/2020\|EPI_ISL_699 | .................... | .................... | .................... | .................... | .................... | .................... |
| W | 2425 | hCoV-19/United Arab Emirates/0952/2020\|EPI_ISL_699 | .................... | .................... | .................... | .................... | .................... | .................... |
| W | 2426 | hCoV-19/United Arab Emirates/0953/2020\|EPI_ISL_699 | .................... | .................... | .................... | .................... | .................... | .................... |
| W | 2427 | hCoV-19/England/ALDP-BB9690/2020\|EPI_ISL_702402\|20 | .................... | .................... | .................... | .................... | .................... | .................... |
| W | 2428 | hCoV-19/England/ALDP-BBB5DD/2020\|EPI_ISL_702404\|20 | .................... | .................... | .................... | .................... | .................... | .................... |
| W | 2429 | hCoV-19/England/ALDP-BBA680/2020\|EPI_ISL_702405\|20 | .................... | .A.................. | .................... | .................... | .................... | ..A................. |
| W | 2430 | hCoV-19/England/ALDP-BBA969/2020\|EPI_ISL_702406\|20 | .................... | .................... | .................... | .................... | .................... | .................... |
| W | 2431 | hCoV-19/England/ALDP-BB92F6/2020\|EPI_ISL_702407\|20 | .................... | .................... | .................... | .................... | .................... | .................... |
| W | 2432 | hCoV-19/England/ALDP-BBADA9/2020\|EPI_ISL_702408\|20 | .................... | .................... | .................... | .................... | .................... | .................... |
| W | 2433 | hCoV-19/England/ALDP-BBA69F/2020\|EPI_ISL_702410\|20 | .................... | .A.................. | .................... | .................... | .................... | ..A................. |
| W | 2434 | hCoV-19/England/ALDP-BB951B/2020\|EPI_ISL_702413\|20 | .................... | .................... | .................... | .................... | .................... | .................... |
| W | 2435 | hCoV-19/England/ALDP-BBADE5/2020\|EPI_ISL_702414\|20 | .................... | .................... | .................... | .................... | .................... | .................... |
| W | 2436 | hCoV-19/England/ALDP-BBA644/2020\|EPI_ISL_702415\|20 | .................... | .A.................. | .................... | .................... | .................... | ..A................. |
| W | 2437 | hCoV-19/England/ALDP-BB9C8F/2020\|EPI_ISL_702417\|20 | .................... | .................... | .................... | .................... | .................... | .................... |
| W | 2438 | hCoV-19/England/ALDP-BB9BA0/2020\|EPI_ISL_702418\|20 | .................... | .................... | .................... | .................... | .................... | .................... |
| W | 2439 | hCoV-19/England/ALDP-BB9681/2020\|EPI_ISL_702419\|20 | .................... | .................... | .................... | .................... | .................... | .................... |
| W | 2440 | hCoV-19/England/ALDP-BB4E51/2020\|EPI_ISL_702422\|20 | .................... | .................... | .................... | .................... | .................... | .................... |
| W | 2441 | hCoV-19/England/ALDP-BB96BE/2020\|EPI_ISL_702423\|20 | .................... | .................... | .................... | .................... | .................... | .................... |
| W | 2442 | hCoV-19/England/ALDP-BBADC7/2020\|EPI_ISL_702424\|20 | .................... | .................... | .................... | .................... | .................... | .................... |
| W | 2443 | hCoV-19/England/ALDP-BBB397/2020\|EPI_ISL_702425\|20 | .................... | .................... | .................... | .................... | .................... | .................... |
| W | 2444 | hCoV-19/England/ALDP-BBA987/2020\|EPI_ISL_702426\|20 | .................... | .................... | .................... | .................... | .................... | .................... |
| W | 2445 | hCoV-19/England/ALDP-BB4D45/2020\|EPI_ISL_702427\|20 | .................... | .................... | .................... | .................... | .................... | .................... |
| W | 2446 | hCoV-19/England/ALDP-BBA6EA/2020\|EPI_ISL_702428\|20 | .................... | .................... | .................... | .................... | .................... | .................... |
| W | 2447 | hCoV-19/England/ALDP-BB9645/2020\|EPI_ISL_702429\|20 | .................... | .A.................. | .................... | .................... | .................... | ..A................. |
| W | 2448 | hCoV-19/England/ALDP-BBA723/2020\|EPI_ISL_702430\|20 | .................... | .A.................. | .................... | .................... | .................... | ..A................. |
| W | 2449 | hCoV-19/England/ALDP-BBAEB5/2020\|EPI_ISL_702431\|20 | .................... | .................... | .................... | .................... | .................... | .................... |
| W | 2450 | hCoV-19/England/ALDP-BBA5A1/2020\|EPI_ISL_702432\|20 | .................... | .................... | .................... | .................... | ......T............. | .................... |
| W | 2451 | hCoV-19/England/ALDP-BB9584/2020\|EPI_ISL_702433\|20 | .................... | .................... | .................... | .................... | .................... | .................... |
| W | 2452 | hCoV-19/England/ALDP-BBA5CF/2020\|EPI_ISL_702434\|20 | .................... | .A.................. | .................... | .................... | .................... | ..A................. |
| W | 2453 | hCoV-19/England/ALDP-BBA95A/2020\|EPI_ISL_702436\|20 | .................... | .................... | .................... | .................... | .................... | .................... |
| W | 2454 | hCoV-19/England/ALDP-BBB564/2020\|EPI_ISL_702437\|20 | .................... | .................... | .................... | .................... | .................... | .................... |
| W | 2455 | hCoV-19/England/ALDP-BB4E06/2020\|EPI_ISL_702439\|20 | .................... | .................... | .................... | .................... | .................... | .................... |
| W | 2456 | hCoV-19/England/ALDP-BBB27C/2020\|EPI_ISL_702440\|20 | .................... | .................... | .................... | .................... | .................... | .................... |
| W | 2457 | hCoV-19/England/ALDP-BBA92D/2020\|EPI_ISL_702441\|20 | .................... | .................... | .................... | .................... | .................... | .................... |
| W | 2458 | hCoV-19/England/ALDP-BBB5EC/2020\|EPI_ISL_702442\|20 | .................... | .................... | .................... | .................... | .................... | .................... |
| W | 2459 | hCoV-19/England/ALDP-BBB591/2020\|EPI_ISL_702443\|20 | .................... | .................... | .................... | .................... | .................... | .................... |
| W | 2460 | hCoV-19/England/ALDP-BBA6BD/2020\|EPI_ISL_702444\|20 | .................... | .................... | .................... | .................... | .................... | .................... |
| W | 2461 | hCoV-19/England/ALDP-BBAAB1/2020\|EPI_ISL_702445\|20 | .................... | .................... | .................... | .................... | .................... | .................... |
| W | 2462 | hCoV-19/England/ALDP-BB4D63/2020\|EPI_ISL_702448\|20 | .................... | .................... | .................... | .................... | .................... | .................... |
| W | 2463 | hCoV-19/England/ALDP-BBB3C4/2020\|EPI_ISL_702449\|20 | .................... | .................... | .................... | .................... | .................... | .................... |
| W | 2464 | hCoV-19/England/ALDP-BBA6DB/2020\|EPI_ISL_702450\|20 | .................... | .................... | .................... | .................... | .................... | .................... |
| W | 2465 | hCoV-19/England/ALDP-BBA9E1/2020\|EPI_ISL_702451\|20 | .................... | .................... | .................... | .................... | .........A.......... | .................... |
| W | 2466 | hCoV-19/England/ALDP-BB9B91/2020\|EPI_ISL_702452\|20 | .................... | .A.................. | .................... | .................... | .................... | ..A................. |
| W | 2467 | hCoV-19/England/ALDP-BB9B82/2020\|EPI_ISL_702454\|20 | .................... | .................... | .................... | .................... | .................... | .................... |
| W | 2468 | hCoV-19/England/ALDP-BBB3D3/2020\|EPI_ISL_702456\|20 | .................... | .................... | .................... | .................... | .................... | .................... |
| W | 2469 | hCoV-19/England/ALDP-BB9320/2020\|EPI_ISL_702457\|20 | .................... | .A.................. | .................... | .................... | .................... | ..A................. |
| W | 2470 | hCoV-19/England/ALDP-BBADD6/2020\|EPI_ISL_702458\|20 | .................... | .................... | .................... | .................... | .................... | .................... |
| W | 2471 | hCoV-19/England/ALDP-BB9C16/2020\|EPI_ISL_702459\|20 | .................... | .................... | .................... | .................... | .................... | .................... |
| W | 2472 | hCoV-19/England/ALDP-BB9C34/2020\|EPI_ISL_702460\|20 | .................... | .................... | .................... | .................... | .................... | .................... |
| W | 2473 | hCoV-19/England/ALDP-BB9548/2020\|EPI_ISL_702461\|20 | .................... | .................... | .................... | .................... | .................... | .................... |
| W | 2474 | hCoV-19/England/ALDP-BBA608/2020\|EPI_ISL_702462\|20 | .................... | .A.................. | .................... | .................... | .................... | ..A................. |
| W | 2475 | hCoV-19/England/ALDP-BBABBE/2020\|EPI_ISL_702463\|20 | .................... | .................... | .................... | .................... | .................... | .................... |
| W | 2476 | hCoV-19/England/ALDP-BBABEB/2020\|EPI_ISL_702465\|20 | .................... | .................... | .................... | .................... | .................... | .................... |
| W | 2477 | hCoV-19/England/ALDP-BB4A3F/2020\|EPI_ISL_702467\|20 | .................... | .................... | .................... | .................... | .................... | .................... |
| W | 2478 | hCoV-19/England/ALDP-BB4A20/2020\|EPI_ISL_702468\|20 | .................... | .................... | .................... | .................... | .................... | .................... |
| W | 2479 | hCoV-19/England/ALDP-BB4E33/2020\|EPI_ISL_702472\|20 | .................... | .................... | .................... | .................... | ......T............. | .................... |
| W | 2480 | hCoV-19/England/ALDP-BBB28B/2020\|EPI_ISL_702475\|20 | .................... | .................... | .................... | .................... | .................... | .................... |
| W | 2481 | hCoV-19/England/ALDP-BBB573/2020\|EPI_ISL_702479\|20 | .................... | .................... | .................... | .................... | .................... | .................... |
| W | 2482 | hCoV-19/England/ALDP-BBB5A0/2020\|EPI_ISL_702480\|20 | .................... | .................... | .................... | .................... | .................... | .................... |
| W | 2483 | hCoV-19/England/ALDP-BB9B73/2020\|EPI_ISL_702482\|20 | .................... | .................... | .................... | .................... | .................... | .................... |
| W | 2484 | hCoV-19/England/ALDP-BB952A/2020\|EPI_ISL_702485\|20 | .................... | .................... | .................... | .................... | .................... | .................... |
| W | 2485 | hCoV-19/England/ALDP-BBB5BF/2020\|EPI_ISL_702486\|20 | .................... | .................... | .................... | .................... | .................... | .................... |
| W | 2486 | hCoV-19/England/ALDP-BBB3A6/2020\|EPI_ISL_702490\|20 | .................... | .................... | .................... | .................... | .................... | .................... |
| W | 2487 | hCoV-19/England/ALDP-BB4DEB/2020\|EPI_ISL_702491\|20 | .................... | .................... | .................... | .................... | .................... | .................... |
| W | 2488 | hCoV-19/England/ALDP-BB4DDC/2020\|EPI_ISL_702493\|20 | .................... | .................... | .................... | .................... | .................... | .................... |

| | | | | | | | | |
|---|---|---|---|---|---|---|---|---|
| W | 2489 | hCoV-19/England/ALDP-BB950C/2020\|EPI_ISL_702494\|20 | ···················· | ···················· | ····················· | ···················· | ···················· | ···················· |
| W | 2490 | hCoV-19/England/ALDP-BB4A5D/2020\|EPI_ISL_702499\|20 | ···················· | ···················· | ····················· | ···················· | ···················· | ···················· |
| W | 2491 | hCoV-19/England/QEUH-BC40FE/2020\|EPI_ISL_703700\|20 | ···················· | ···················· | ····················· | ···················· | ···················· | ···················· |
| W | 2492 | hCoV-19/England/ALDP-BCAA81/2020\|EPI_ISL_703702\|20 | ···················· | ···················· | ····················· | ···················· | ···················· | ···················· |
| W | 2493 | hCoV-19/England/QEUH-BC400D/2020\|EPI_ISL_703703\|20 | ···················· | ···················· | ····················· | ···················· | ···················· | ···················· |
| W | 2494 | hCoV-19/England/ALDP-BCA9EE/2020\|EPI_ISL_703705\|20 | ···················· | ···················· | ····················· | ···················· | ···················· | ···A················ |
| W | 2495 | hCoV-19/England/ALDP-BCA878/2020\|EPI_ISL_703707\|20 | ···················· | ···················· | ····················· | ···················· | ···················· | ···················· |
| W | 2496 | hCoV-19/England/ALDP-BCA535/2020\|EPI_ISL_703777\|20 | ···················· | ···················· | ····················· | ···················· | ···················· | ···················· |
| W | 2497 | hCoV-19/England/ALDP-BCA474/2020\|EPI_ISL_703778\|20 | ···················· | ···················· | ····················· | ···················· | ···················· | ···················· |
| W | 2498 | hCoV-19/England/ALDP-BCA24D/2020\|EPI_ISL_703780\|20 | ···················· | ···················· | ····················· | ···················· | ···················· | ···················· |
| W | 2499 | hCoV-19/England/ALDP-BCA711/2020\|EPI_ISL_703781\|20 | ···················· | ···················· | ····················· | ···················· | ···················· | ···················· |
| W | 2500 | hCoV-19/England/ALDP-BCA90C/2020\|EPI_ISL_703783\|20 | ···················· | ···················· | ····················· | ···················· | ···················· | ···················· |
| W | 2501 | hCoV-19/England/QEUH-BC4559/2020\|EPI_ISL_703786\|20 | ···················· | ···················· | ····················· | ···················· | ···················· | ···················· |
| W | 2502 | hCoV-19/England/ALDP-BCA17D/2020\|EPI_ISL_703788\|20 | ···················· | ···················· | ····················· | ···················· | ···················· | ···················· |
| W | 2503 | hCoV-19/England/QEUH-BC4252/2020\|EPI_ISL_703790\|20 | ···················· | ···················· | ····················· | ···················· | ···················· | ···················· |
| W | 2504 | hCoV-19/England/ALDP-BCAB7F/2020\|EPI_ISL_703791\|20 | ···················· | ···················· | ····················· | ···················· | ···················· | ···················· |
| W | 2505 | hCoV-19/England/ALDP-BCA2E3/2020\|EPI_ISL_703793\|20 | ···················· | ···················· | ····················· | ···················· | ···················· | ···················· |
| W | 2506 | hCoV-19/England/ALDP-BCA395/2020\|EPI_ISL_703795\|20 | ···················· | ···················· | ····················· | ···················· | ···················· | ···················· |
| W | 2507 | hCoV-19/England/ALDP-BCA74E/2020\|EPI_ISL_703796\|20 | ···················· | ···················· | ····················· | ···················· | ···················· | ···················· |
| W | 2508 | hCoV-19/England/ALDP-BCA7F3/2020\|EPI_ISL_703798\|20 | ···················· | ···················· | ····················· | ···················· | ···················· | ···················· |
| W | 2509 | hCoV-19/England/QEUH-BC4805/2020\|EPI_ISL_703799\|20 | ···················· | ···················· | ····················· | ···················· | ···················· | ···················· |
| W | 2510 | hCoV-19/USA/MI-MDHHS-SC22541/2020\|EPI_ISL_708600\|2 | ···················· | ···················· | ····················· | ···················· | ···················· | ···················· |
| W | 2511 | hCoV-19/USA/MI-MDHHS-SC22542/2020\|EPI_ISL_708601\|2 | ···················· | ·············A······ | ····················· | ···················· | ···················· | ···················· |
| W | 2512 | hCoV-19/USA/MI-MDHHS-SC22543/2020\|EPI_ISL_708602\|2 | ···················· | ···················· | ····················· | ···················· | ···················· | ···················· |
| W | 2513 | hCoV-19/USA/MI-MDHHS-SC22545/2020\|EPI_ISL_708603\|2 | ···················· | ···················· | ····················· | ···················· | ···················· | ···················· |
| W | 2514 | hCoV-19/USA/MI-MDHHS-SC22546/2020\|EPI_ISL_708604\|2 | ···················· | ···················· | ····················· | ···················· | ···················· | ···················· |
| W | 2515 | hCoV-19/USA/MI-MDHHS-SC22547/2020\|EPI_ISL_708605\|2 | ···················· | ···················· | ····················· | ···················· | ···················· | ···················· |
| W | 2516 | hCoV-19/USA/MI-MDHHS-SC22548/2020\|EPI_ISL_708606\|2 | ···················· | ···················· | ····················· | ···················· | ···················· | ···················· |
| W | 2517 | hCoV-19/USA/TX-DSHS-1726/2020\|EPI_ISL_708650\|2020- | ···················· | ···················· | ····················· | ···················· | ···················· | ···················· |
| W | 2518 | hCoV-19/Indonesia/BT-SHSIU-01-4920/2020\|EPI_ISL_70 | ···················· | ····A··············· | ····················· | ···················· | ···················· | ···················· |
| W | 2519 | hCoV-19/England/MILK-B5506B/2020\|EPI_ISL_709900\|20 | ···················· | ···················· | ····················· | ···················· | ···················· | ···················· |
| W | 2520 | hCoV-19/England/MILK-B55256/2020\|EPI_ISL_709901\|20 | ···················· | ···················· | ····················· | ···················· | ···················· | ···················· |
| W | 2521 | hCoV-19/England/MILK-B56088/2020\|EPI_ISL_709902\|20 | ···················· | ·A·················· | ····················· | ···················· | ···················· | ···A················ |
| W | 2522 | hCoV-19/England/MILK-B38D94/2020\|EPI_ISL_709903\|20 | ···················· | ···················· | ····················· | ···················· | ···················· | ···················· |
| W | 2523 | hCoV-19/England/MILK-B38DB2/2020\|EPI_ISL_709904\|20 | ···················· | ···················· | ····················· | ···················· | ···················· | ···················· |
| W | 2524 | hCoV-19/England/MILK-B3830B/2020\|EPI_ISL_709905\|20 | ···················· | ···················· | ····················· | ···················· | ···················· | ···················· |
| W | 2525 | hCoV-19/England/MILK-B02777/2020\|EPI_ISL_709906\|20 | ···················· | ···················· | ····················· | ···················· | ···················· | ···················· |
| W | 2526 | hCoV-19/England/MILK-B027B3/2020\|EPI_ISL_709907\|20 | ···················· | ···················· | ····················· | ···················· | ···················· | ···················· |
| W | 2527 | hCoV-19/England/MILK-B0272C/2020\|EPI_ISL_709908\|20 | ···················· | ···················· | ····················· | ···················· | ···················· | ···················· |
| W | 2528 | hCoV-19/England/MILK-B022D0/2020\|EPI_ISL_709909\|20 | ···················· | ···················· | ····················· | ···················· | ···················· | ···················· |
| W | 2529 | hCoV-19/England/MILK-B021F1/2020\|EPI_ISL_709910\|20 | ···················· | ·A·················· | ····················· | ···················· | ···················· | ···A················ |
| W | 2530 | hCoV-19/England/MILK-ACFFBB/2020\|EPI_ISL_709911\|20 | ···················· | ···················· | ····················· | ···················· | ···················· | ···················· |
| W | 2531 | hCoV-19/England/MILK-AD00D2/2020\|EPI_ISL_709912\|20 | ···················· | ···················· | ····················· | ···················· | ···················· | ···················· |
| W | 2532 | hCoV-19/England/MILK-ACFFD9/2020\|EPI_ISL_709913\|20 | ···················· | ·A·················· | ····················· | ···················· | ···················· | ···A················ |
| W | 2533 | hCoV-19/England/MILK-AC9142/2020\|EPI_ISL_709920\|20 | ···················· | ···················· | ····················· | ···················· | ···················· | ···················· |
| W | 2534 | hCoV-19/England/MILK-AC9160/2020\|EPI_ISL_709921\|20 | ···················· | ···················· | ····················· | ···················· | ···················· | ···················· |
| W | 2535 | hCoV-19/England/MILK-AC9300/2020\|EPI_ISL_709922\|20 | ···················· | ···················· | ····················· | ···················· | ···················· | ···················· |
| W | 2536 | hCoV-19/England/MILK-AC8FC1/2020\|EPI_ISL_709923\|20 | ···················· | ···················· | ····················· | ···················· | ······T············· | ···············A···· |
| W | 2537 | hCoV-19/England/MILK-AC9573/2020\|EPI_ISL_709924\|20 | ···················· | ···················· | ····················· | ···················· | ···················· | ···················· |
| W | 2538 | hCoV-19/England/MILK-AB79B8/2020\|EPI_ISL_709925\|20 | ···················· | ···················· | ····················· | ···················· | ···················· | ···················· |
| W | 2539 | hCoV-19/England/MILK-AB7C0A/2020\|EPI_ISL_709927\|20 | ···················· | ···················· | ····················· | ···················· | ···················· | ···················· |
| W | 2540 | hCoV-19/England/MILK-AB7860/2020\|EPI_ISL_709928\|20 | ···················· | ···················· | ····················· | ···················· | ······T············· | ···················· |
| W | 2541 | hCoV-19/England/MILK-AB787F/2020\|EPI_ISL_709931\|20 | ···················· | ···················· | ····················· | ···················· | ······T············· | ···················· |
| W | 2542 | hCoV-19/England/MILK-A7D8B1/2020\|EPI_ISL_709932\|20 | ···················· | ···················· | ····················· | ···················· | ···················· | ···················· |
| W | 2543 | hCoV-19/England/MILK-A7D75A/2020\|EPI_ISL_709933\|20 | ···················· | ···················· | ····················· | ···················· | ···················· | ···················· |
| W | 2544 | hCoV-19/England/MILK-A7DF25/2020\|EPI_ISL_709935\|20 | ···················· | ···················· | ····················· | ···················· | ···················· | ···················· |
| W | 2545 | hCoV-19/England/MILK-A7A5F9/2020\|EPI_ISL_709936\|20 | ···················· | ···················· | ····················· | ···················· | ···················· | ···················· |
| W | 2546 | hCoV-19/England/MILK-A797F4/2020\|EPI_ISL_709938\|20 | ···················· | ···················· | ····················· | ···················· | ···················· | ···················· |
| W | 2547 | hCoV-19/England/MILK-A7983D/2020\|EPI_ISL_709939\|20 | ···················· | ···················· | ····················· | ···················· | ···················· | ···················· |
| W | 2548 | hCoV-19/England/MILK-A7981F/2020\|EPI_ISL_709940\|20 | ···················· | ···················· | ····················· | ···················· | ···················· | ···················· |
| W | 2549 | hCoV-19/England/MILK-A7A15F/2020\|EPI_ISL_709941\|20 | ···················· | ·A·················· | ····················· | ···················· | ···················· | ···A················ |
| W | 2550 | hCoV-19/Wales/MILK-A6B704/2020\|EPI_ISL_709942\|2020 | ···················· | ···················· | ····················· | ···················· | ···················· | ···················· |
| W | 2551 | hCoV-19/England/MILK-A6BA1A/2020\|EPI_ISL_709943\|20 | ···················· | ···················· | ····················· | ···················· | ···················· | ···················· |
| W | 2552 | hCoV-19/England/MILK-A6AB54/2020\|EPI_ISL_709944\|20 | ···················· | ···················· | ····················· | ···················· | ···················· | ···················· |
| W | 2553 | hCoV-19/England/MILK-A6B86B/2020\|EPI_ISL_709945\|20 | ···················· | ·A·················· | ····················· | ···················· | ···················· | ···················· |
| W | 2554 | hCoV-19/England/205000526/2020\|EPI_ISL_709958\|2020 | ···················· | ···················· | ····················· | ···················· | ···················· | ···················· |
| W | 2555 | hCoV-19/Gibraltar/205000623/2020\|EPI_ISL_709959\|20 | ···················· | ···················· | ····················· | ···················· | ···················· | ···················· |
| W | 2556 | hCoV-19/Gibraltar/205000645/2020\|EPI_ISL_709980\|20 | ···················· | ···················· | ····················· | ···················· | ···················· | ···················· |
| W | 2557 | hCoV-19/Gibraltar/205000650/2020\|EPI_ISL_709985\|20 | ···················· | ···················· | ····················· | ···················· | ···················· | ···················· |
| W | 2558 | hCoV-19/Gibraltar/205000651/2020\|EPI_ISL_709986\|20 | ···················· | ···················· | ····················· | ···················· | ···················· | ···················· |
| W | 2559 | hCoV-19/Gibraltar/205000653/2020\|EPI_ISL_709987\|20 | ···················· | ···················· | ····················· | ···················· | ···················· | ···················· |
| W | 2560 | hCoV-19/Gibraltar/205000657/2020\|EPI_ISL_709988\|20 | ···················· | ···················· | ····················· | ···················· | ···················· | ···················· |
| W | 2561 | hCoV-19/USA/AZ-TG276187/2020\|EPI_ISL_694100\|2020-0 | ···················· | ···················· | ····················· | ···················· | ···················· | ···················· |
| W | 2562 | hCoV-19/USA/AZ-TG276456/2020\|EPI_ISL_694101\|2020-0 | ···················· | ···················· | ····················· | ···················· | ···················· | ···················· |
| W | 2563 | hCoV-19/USA/AZ-TG276458/2020\|EPI_ISL_694102\|2020-0 | ···················· | ···················· | ····················· | ···················· | ···················· | ···················· |
| W | 2564 | hCoV-19/USA/AZ-TG276460/2020\|EPI_ISL_694103\|2020-0 | ···················· | ···················· | ····················· | ···················· | ···················· | ···················· |
| W | 2565 | hCoV-19/USA/AZ-TG276462/2020\|EPI_ISL_694104\|2020-0 | ···················· | ···················· | ····················· | ···················· | ···················· | ···················· |
| W | 2566 | hCoV-19/USA/AZ-TG276476/2020\|EPI_ISL_694105\|2020-0 | ···················· | ···················· | ····················· | ···················· | ···················· | ···················· |
| W | 2567 | hCoV-19/USA/AZ-TG276490/2020\|EPI_ISL_694106\|2020-0 | ···················· | ···················· | ····················· | ···················· | ···················· | ···················· |
| W | 2568 | hCoV-19/USA/AZ-TG276492/2020\|EPI_ISL_694107\|2020-0 | ···················· | ···················· | ····················· | ···················· | ···················· | ···················· |

| | | | | | | | |
|---|---|---|---|---|---|---|---|
| W | 2569 hCoV-19/USA/AZ-TG276577/2020\|EPI_ISL_694112\|2020-0 | ···················· | ···················· | ···················· | ···················· | ···················· | ···················· |
| W | 2570 hCoV-19/USA/AZ-TG280820/2020\|EPI_ISL_694166\|2020-0 | ···················· | ···················· | ···················· | ···················· | ···················· | ···················· |
| W | 2571 hCoV-19/USA/AZ-TG280821/2020\|EPI_ISL_694167\|2020-0 | ···················· | ···················· | ···················· | ···················· | ···················· | ···················· |
| W | 2572 hCoV-19/USA/AZ-TG280822/2020\|EPI_ISL_694168\|2020-0 | ···················· | ···················· | ···················· | ···················· | ···················· | ···················· |
| W | 2573 hCoV-19/USA/AZ-TG280830/2020\|EPI_ISL_694173\|2020-0 | ···················· | ···················· | ···················· | ···················· | ···················· | ···················· |
| W | 2574 hCoV-19/USA/AZ-TG280831/2020\|EPI_ISL_694174\|2020-0 | ···················· | ···················· | ···················· | ···················· | ···················· | ···················· |
| W | 2575 hCoV-19/USA/AZ-TG280834/2020\|EPI_ISL_694177\|2020-0 | ···················· | ···················· | ···················· | ···················· | ···················· | ···················· |
| W | 2576 hCoV-19/USA/AZ-TG280845/2020\|EPI_ISL_694186\|2020-0 | ···················· | ···················· | ···················· | ···················· | ···················· | ···················· |
| W | 2577 hCoV-19/USA/AZ-TG280846/2020\|EPI_ISL_694187\|2020-0 | ···················· | ···················· | ···················· | ···················· | ···················· | ···················· |
| W | 2578 hCoV-19/USA/AZ-TG280848/2020\|EPI_ISL_694189\|2020-0 | ···················· | ···················· | ···················· | ···················· | ···················· | ···················· |
| W | 2579 hCoV-19/USA/AZ-TG280859/2020\|EPI_ISL_694198\|2020-0 | ···················· | ···················· | ···················· | ···················· | ···················· | ···················· |
| W | 2580 hCoV-19/USA/AZ-TG280865/2020\|EPI_ISL_694199\|2020-0 | ···················· | ···················· | ···················· | ···················· | ···················· | ···················· |
| W | 2582 hCoV-19/USA/AZ-TG532076/2020\|EPI_ISL_695406\|2020-0 | ···················· | ···················· | ···················· | ···················· | ···················· | ···················· |
| W | 2583 hCoV-19/USA/AZ-TG532081/2020\|EPI_ISL_695408\|2020-0 | ···················· | ···················· | ···················· | ···················· | ···················· | ···················· |
| W | 2584 hCoV-19/USA/AZ-TG532086/2020\|EPI_ISL_695411\|2020-0 | ···················· | ···················· | ···················· | ···················· | ···················· | ···················· |
| W | 2586 hCoV-19/USA/AZ-TG532098/2020\|EPI_ISL_695415\|2020-0 | ···················· | ···················· | ···················· | ···················· | ···················· | ···················· |
| W | 2587 hCoV-19/USA/AZ-TG532112/2020\|EPI_ISL_695419\|2020-0 | ···················· | ·A·················· | ···················· | ···················· | ···················· | ···················· |
| W | 2588 hCoV-19/USA/AZ-TG532122/2020\|EPI_ISL_695423\|2020-0 | ···················· | ·A·················· | ···················· | ···················· | ···················· | ···················· |
| W | 2589 hCoV-19/USA/AZ-TG547214/2020\|EPI_ISL_695426\|2020-0 | ···················· | ···················· | ···················· | ···················· | ···················· | ···················· |
| W | 2590 hCoV-19/USA/AZ-TG547215/2020\|EPI_ISL_695427\|2020-0 | ···················· | ···················· | ···················· | ···················· | ···················· | ···················· |
| W | 2591 hCoV-19/USA/AZ-TG547216/2020\|EPI_ISL_695428\|2020-0 | ···················· | ···················· | ···················· | ···················· | ···················· | ···················· |
| W | 2592 hCoV-19/USA/AZ-TG547217/2020\|EPI_ISL_695429\|2020-0 | ···················· | ···················· | ···················· | ···················· | ···················· | ···················· |
| W | 2593 hCoV-19/USA/AZ-TG547220/2020\|EPI_ISL_695431\|2020-0 | ···················· | ···················· | ···················· | ···················· | ···················· | ···················· |
| W | 2594 hCoV-19/USA/AZ-TG547222/2020\|EPI_ISL_695433\|2020-0 | ···················· | ············A······· | ···················· | ···················· | ···················· | ···················· |
| W | 2595 hCoV-19/USA/AZ-TG547223/2020\|EPI_ISL_695434\|2020-0 | ···················· | ············A······· | ···················· | ···················· | ···················· | ···················· |
| W | 2596 hCoV-19/USA/AZ-TG547225/2020\|EPI_ISL_695435\|2020-0 | ···················· | ·A·················· | ···················· | ···················· | ···················· | ···················· |
| W | 2597 hCoV-19/USA/AZ-TG547230/2020\|EPI_ISL_695437\|2020-0 | ···················· | ···················· | ···················· | ···················· | ···················· | ···················· |
| W | 2598 hCoV-19/USA/AZ-TG547231/2020\|EPI_ISL_695438\|2020-0 | ···················· | ···················· | ···················· | ···················· | ···················· | ···················· |
| W | 2599 hCoV-19/USA/AZ-TG547233/2020\|EPI_ISL_695440\|2020-0 | ···················· | ···················· | ···················· | ···················· | ···················· | ···················· |
| W | 2600 hCoV-19/USA/AZ-TG547243/2020\|EPI_ISL_695442\|2020-0 | ···················· | ···················· | ···················· | ···················· | ···················· | ···················· |
| W | 2601 hCoV-19/USA/AZ-TG547343/2020\|EPI_ISL_695491\|2020-0 | ···················· | ·A·················· | ···················· | ···················· | ···················· | ···················· |
| W | 2602 hCoV-19/USA/AZ-TG547344/2020\|EPI_ISL_695492\|2020-0 | ···················· | ·A·················· | ···················· | ···················· | ···················· | ···················· |
| W | 2603 hCoV-19/USA/AZ-TG547345/2020\|EPI_ISL_695493\|2020-0 | ···················· | ·A·················· | ···················· | ···················· | ···················· | ···················· |
| W | 2604 hCoV-19/USA/AZ-TG547346/2020\|EPI_ISL_695494\|2020-0 | ···················· | ·A·················· | ···················· | ···················· | ···················· | ···················· |
| W | 2605 hCoV-19/England/ALDP-BB92E7/2020\|EPI_ISL_702501\|20 | ···················· | ···················· | ···················· | ···················· | ···················· | ···················· |
| W | 2606 hCoV-19/England/ALDP-BB9BEC/2020\|EPI_ISL_702502\|20 | ···················· | ···················· | ···················· | ···················· | ···················· | ···················· |
| W | 2607 hCoV-19/England/ALDP-BB4E15/2020\|EPI_ISL_702503\|20 | ···················· | ···················· | ···················· | ···················· | ···················· | ···················· |
| W | 2608 hCoV-19/England/ALDP-BBB607/2020\|EPI_ISL_702506\|20 | ···················· | ···················· | ···················· | ···················· | ···················· | ···················· |
| W | 2609 hCoV-19/England/ALDP-BB4D81/2020\|EPI_ISL_702509\|20 | ···················· | ···················· | ···················· | ···················· | ···················· | ···················· |
| W | 2610 hCoV-19/England/ALDP-BBA671/2020\|EPI_ISL_702511\|20 | ···················· | ···················· | ···················· | ···················· | ···················· | ···················· |
| W | 2611 hCoV-19/England/ALDP-BB4D54/2020\|EPI_ISL_702514\|20 | ···················· | ···················· | ···················· | ···················· | ···················· | ···················· |
| W | 2612 hCoV-19/England/ALDP-BBA996/2020\|EPI_ISL_702516\|20 | ···················· | ···················· | ···················· | ···················· | ···················· | ···················· |
| W | 2613 hCoV-19/England/ALDP-BBA592/2020\|EPI_ISL_702517\|20 | ···················· | ···················· | ···················· | ···················· | ·····T·············· | ···················· |
| W | 2614 hCoV-19/England/ALDP-BB47CF/2020\|EPI_ISL_702519\|20 | ···················· | ···················· | ···················· | ···················· | ···················· | ···················· |
| W | 2615 hCoV-19/England/ALDP-BBAD9A/2020\|EPI_ISL_702521\|20 | ···················· | ···················· | ···················· | ···················· | ···················· | ···················· |
| W | 2616 hCoV-19/England/ALDP-BB4ECA/2020\|EPI_ISL_702522\|20 | ···················· | ···················· | ···················· | ···················· | ···················· | ···················· |
| W | 2617 hCoV-19/England/ALDP-BB9557/2020\|EPI_ISL_702524\|20 | ···················· | ···················· | ···················· | ···················· | ···················· | ···················· |
| W | 2618 hCoV-19/England/ALDP-BB9C70/2020\|EPI_ISL_702526\|20 | ···················· | ···················· | ···················· | ···················· | ···················· | ···················· |
| W | 2619 hCoV-19/England/ALDP-BBA9A5/2020\|EPI_ISL_702527\|20 | ···················· | ···················· | ···················· | ···················· | ···················· | ···················· |
| W | 2620 hCoV-19/England/ALDP-BBB3B5/2020\|EPI_ISL_702529\|20 | ···················· | ···················· | ···················· | ···················· | ···················· | ···················· |
| W | 2621 hCoV-19/England/ALDP-BBB40D/2020\|EPI_ISL_702530\|20 | ···················· | ···················· | ···················· | ···················· | ···················· | ···················· |
| W | 2622 hCoV-19/England/ALDP-BBAA1B/2020\|EPI_ISL_702532\|20 | ···················· | ···················· | ···················· | ···················· | ···················· | ···················· |
| W | 2623 hCoV-19/England/ALDP-BB96CD/2020\|EPI_ISL_702534\|20 | ···················· | ···················· | ···················· | ···················· | ···················· | ···················· |
| W | 2624 hCoV-19/England/ALDP-BBB537/2020\|EPI_ISL_702535\|20 | ···················· | ···················· | ···················· | ···················· | ···················· | ···················· |
| W | 2625 hCoV-19/England/ALDP-BBAD6D/2020\|EPI_ISL_702537\|20 | ···················· | ···················· | ···················· | ···················· | ·····T·············· | ···················· |
| W | 2626 hCoV-19/England/ALDP-BBABDC/2020\|EPI_ISL_702538\|20 | ···················· | ···················· | ···················· | ···················· | ···················· | ···················· |
| W | 2627 hCoV-19/England/ALDP-BB92BA/2020\|EPI_ISL_702542\|20 | ···················· | ···················· | ···················· | ···················· | ···················· | ···················· |
| W | 2628 hCoV-19/England/ALDP-BBAC06/2020\|EPI_ISL_702546\|20 | ···················· | ···················· | ···················· | ···················· | ···················· | ···················· |
| W | 2629 hCoV-19/England/ALDP-BBAA93/2020\|EPI_ISL_702555\|20 | ···················· | ···················· | ···················· | ···················· | ···················· | ···················· |
| W | 2630 hCoV-19/England/ALDP-BB4D90/2020\|EPI_ISL_702557\|20 | ···················· | ···················· | ···················· | ···················· | ···················· | ···················· |
| W | 2631 hCoV-19/England/ALDP-BBB519/2020\|EPI_ISL_702559\|20 | ···················· | ···················· | ···················· | ···················· | ···················· | ···················· |
| W | 2632 hCoV-19/England/ALDP-BB4941/2020\|EPI_ISL_702563\|20 | ···················· | ···················· | ···················· | ···················· | ···················· | ···················· |
| W | 2633 hCoV-19/England/ALDP-BBA5DE/2020\|EPI_ISL_702565\|20 | ···················· | ···················· | ···················· | ···················· | ···················· | ···················· |
| W | 2634 hCoV-19/England/ALDP-BB9302/2020\|EPI_ISL_702567\|20 | ···················· | ···················· | ···················· | ···················· | ···················· | ···················· |
| W | 2635 hCoV-19/England/ALDP-BBB582/2020\|EPI_ISL_702568\|20 | ···················· | ···················· | ···················· | ···················· | ···················· | ···················· |
| W | 2636 hCoV-19/England/QEUH-BC3F98/2020\|EPI_ISL_703801\|20 | ···················· | ···················· | ···················· | ···················· | ···················· | ···················· |
| W | 2637 hCoV-19/England/ALDP-BCA3E0/2020\|EPI_ISL_703803\|20 | ···················· | ···················· | ···················· | ···················· | ···················· | ···················· |
| W | 2638 hCoV-19/England/ALDP-BCA438/2020\|EPI_ISL_703804\|20 | ···················· | ···················· | ···················· | ···················· | ···················· | ···················· |
| W | 2639 hCoV-19/England/QEUH-BC487E/2020\|EPI_ISL_703808\|20 | ···················· | ···················· | ···················· | ···················· | ···················· | ···················· |
| W | 2640 hCoV-19/England/ALDP-BCAADC/2020\|EPI_ISL_703809\|20 | ···················· | ···················· | ···················· | ···················· | ···················· | ···················· |
| W | 2641 hCoV-19/England/ALDP-BCA73F/2020\|EPI_ISL_703812\|20 | ···················· | ···················· | ···················· | ···················· | ···················· | ···················· |
| W | 2642 hCoV-19/England/ALDP-BCA8B4/2020\|EPI_ISL_703814\|20 | ···················· | ···················· | ···················· | ···················· | ···················· | ···················· |
| W | 2643 hCoV-19/England/QEUH-BC4665/2020\|EPI_ISL_703816\|20 | ···················· | ···················· | ···················· | ···················· | ···················· | ···················· |
| W | 2644 hCoV-19/England/ALDP-BCA4FC/2020\|EPI_ISL_703817\|20 | ···················· | ···················· | ···················· | ···················· | ···················· | ···················· |
| W | 2645 hCoV-19/England/QEUH-BC4B84/2020\|EPI_ISL_703819\|20 | ···················· | ···················· | ···················· | ···················· | ···················· | ···················· |
| W | 2646 hCoV-19/England/QEUH-BCC032/2020\|EPI_ISL_703820\|20 | ···················· | ···················· | ···················· | ···················· | ···················· | ···················· |
| W | 2647 hCoV-19/England/QEUH-BCB7F2/2020\|EPI_ISL_703822\|20 | ···················· | ···················· | ···················· | ···················· | ···················· | ···················· |
| W | 2648 hCoV-19/England/QEUH-BCBA17/2020\|EPI_ISL_703824\|20 | ···················· | ···················· | ···················· | ···················· | ···················· | ···················· |
| W | 2649 hCoV-19/England/QEUH-BC3A88/2020\|EPI_ISL_703825\|20 | ···················· | ···················· | ···················· | ···················· | ···················· | ···················· |
| W | 2650 hCoV-19/England/QEUH-BC3A6A/2020\|EPI_ISL_703827\|20 | ···················· | ···················· | ···················· | ···················· | ···················· | ···················· |

| | | | | | | | | |
|---|---|---|---|---|---|---|---|---|
| W | 2651 | hCoV-19/England/QEUH-BCB6AA/2020\|EPI_ISL_703830\|20 | ···················· | ···················· | ····················· | ···················· | ···················· | ···················· |
| W | 2652 | hCoV-19/England/QEUH-BCC7A6/2020\|EPI_ISL_703835\|20 | ···················· | ···················· | ····················· | ···················· | ···················· | ···················· |
| W | 2653 | hCoV-19/England/QEUH-BCB947/2020\|EPI_ISL_703836\|20 | ···················· | ···················· | ····················· | ···················· | ···················· | ···················· |
| W | 2654 | hCoV-19/England/QEUH-BC3781/2020\|EPI_ISL_703838\|20 | ···················· | ···················· | ····················· | ···················· | ···················· | ···················· |
| W | 2655 | hCoV-19/England/QEUH-BC49B7/2020\|EPI_ISL_703840\|20 | ···················· | ···················· | ····················· | ···················· | ···················· | ···················· |
| W | 2656 | hCoV-19/England/QEUH-BC3CA0/2020\|EPI_ISL_703841\|20 | ···················· | ···················· | ····················· | ···················· | ···················· | ···················· |
| W | 2657 | hCoV-19/England/QEUH-BC4A96/2020\|EPI_ISL_703843\|20 | ···················· | ···················· | ····················· | ···················· | ······T············· | ···················· |
| W | 2658 | hCoV-19/England/QEUH-BC3842/2020\|EPI_ISL_703849\|20 | ···················· | ···················· | ····················· | ···················· | ···················· | ···················· |
| W | 2659 | hCoV-19/England/QEUH-BC49F3/2020\|EPI_ISL_703851\|20 | ···················· | ···················· | ····················· | ···················· | ···················· | ···················· |
| W | 2660 | hCoV-19/England/QEUH-BC3E04/2020\|EPI_ISL_703852\|20 | ···················· | ···················· | ····················· | ···················· | ···················· | ···················· |
| W | 2661 | hCoV-19/England/QEUH-BCB965/2020\|EPI_ISL_703854\|20 | ···················· | ···················· | ····················· | ···················· | ···················· | ···················· |
| W | 2662 | hCoV-19/England/QEUH-BC4BB1/2020\|EPI_ISL_703857\|20 | ···················· | ···················· | ····················· | ···················· | ···················· | ···················· |
| W | 2663 | hCoV-19/England/QEUH-BCB82C/2020\|EPI_ISL_703861\|20 | ···················· | ···················· | ····················· | ···················· | ···················· | ···················· |
| W | 2664 | hCoV-19/England/QEUH-BCB65F/2020\|EPI_ISL_703862\|20 | ···················· | ···················· | ····················· | ···················· | ···················· | ···················· |
| W | 2665 | hCoV-19/England/QEUH-BCBD69/2020\|EPI_ISL_703864\|20 | ···················· | ···················· | ····················· | ···················· | ···················· | ···················· |
| W | 2666 | hCoV-19/England/QEUH-BC498A/2020\|EPI_ISL_703866\|20 | ···················· | ···················· | ····················· | ···················· | ···················· | ···················· |
| W | 2667 | hCoV-19/England/QEUH-BC36C0/2020\|EPI_ISL_703869\|20 | ···················· | ···················· | ····················· | ···················· | ···················· | ···················· |
| W | 2668 | hCoV-19/England/QEUH-BC3693/2020\|EPI_ISL_703878\|20 | ···················· | ···················· | ····················· | ···················· | ···················· | ···················· |
| W | 2669 | hCoV-19/England/QEUH-BC4BA2/2020\|EPI_ISL_703880\|20 | ···················· | ···················· | ····················· | ···················· | ······T············· | ···················· |
| W | 2670 | hCoV-19/England/QEUH-BC4B57/2020\|EPI_ISL_703882\|20 | ···················· | ···················· | ····················· | ···················· | ···················· | ···················· |
| W | 2671 | hCoV-19/England/QEUH-BCBCD5/2020\|EPI_ISL_703883\|20 | ···················· | ···················· | ····················· | ···················· | ···················· | ···················· |
| W | 2672 | hCoV-19/England/QEUH-BC3824/2020\|EPI_ISL_703885\|20 | ···················· | ···················· | ····················· | ···················· | ···················· | ···················· |
| W | 2673 | hCoV-19/England/QEUH-BC3C46/2020\|EPI_ISL_703887\|20 | ···················· | ·A·················· | ····················· | ···················· | ···················· | ···················· |
| W | 2674 | hCoV-19/England/QEUH-BC3D34/2020\|EPI_ISL_703889\|20 | ···················· | ···················· | ····················· | ···················· | ···················· | ···················· |
| W | 2675 | hCoV-19/England/QEUH-BC3A4C/2020\|EPI_ISL_703890\|20 | ···················· | ···················· | ····················· | ···················· | ···················· | ···················· |
| W | 2676 | hCoV-19/England/QEUH-BC3E6E/2020\|EPI_ISL_703893\|20 | ···················· | ···················· | ····················· | ···················· | ···················· | ···················· |
| W | 2677 | hCoV-19/England/QEUH-BC3BFE/2020\|EPI_ISL_703895\|20 | ···················· | ·A·················· | ····················· | ···················· | ···················· | ···················· |
| W | 2678 | hCoV-19/England/MILK-A919C3/2020\|EPI_ISL_706000\|20 | ···················· | ···················· | ····················· | ···················· | ···················· | ···················· |
| W | 2679 | hCoV-19/England/MILK-A919F0/2020\|EPI_ISL_706001\|20 | ···················· | ···················· | ····················· | ···················· | ···················· | ···················· |
| W | 2680 | hCoV-19/England/MILK-A91A0C/2020\|EPI_ISL_706002\|20 | ···················· | ·A·················· | ····················· | ···················· | ···················· | ···A················ |
| W | 2681 | hCoV-19/England/MILK-A91A1B/2020\|EPI_ISL_706003\|20 | ···················· | ···················· | ····················· | ···················· | ···················· | ···················· |
| W | 2682 | hCoV-19/England/MILK-A91A2A/2020\|EPI_ISL_706004\|20 | ···················· | ···················· | ····················· | ···················· | ···················· | ···················· |
| W | 2683 | hCoV-19/England/MILK-A91A39/2020\|EPI_ISL_706005\|20 | ···················· | ···················· | ····················· | ···················· | ···················· | ···················· |
| W | 2684 | hCoV-19/England/MILK-A91A75/2020\|EPI_ISL_706006\|20 | ···················· | ···················· | ····················· | ···················· | ···················· | ···················· |
| W | 2685 | hCoV-19/England/MILK-A91AA2/2020\|EPI_ISL_706007\|20 | ···················· | ···················· | ····················· | ···················· | ···················· | ···················· |
| W | 2686 | hCoV-19/England/MILK-A91AB1/2020\|EPI_ISL_706008\|20 | ···················· | ···················· | ····················· | ···················· | ···················· | ···················· |
| W | 2687 | hCoV-19/England/MILK-A91AC0/2020\|EPI_ISL_706009\|20 | ···················· | ···················· | ····················· | ···················· | ···················· | ···················· |
| W | 2688 | hCoV-19/England/MILK-A91ADF/2020\|EPI_ISL_706010\|20 | ···················· | ···················· | ····················· | ···················· | ···················· | ···················· |
| W | 2689 | hCoV-19/England/MILK-A91AFD/2020\|EPI_ISL_706011\|20 | ···················· | ···················· | ····················· | ···················· | ···················· | ···················· |
| W | 2690 | hCoV-19/England/MILK-A91B81/2020\|EPI_ISL_706012\|20 | ···················· | ···················· | ····················· | ···················· | ···················· | ···················· |
| W | 2691 | hCoV-19/England/MILK-B39FAC/2020\|EPI_ISL_706013\|20 | ···················· | ···················· | ····················· | ···················· | ···················· | ···················· |
| W | 2692 | hCoV-19/England/MILK-B3A06C/2020\|EPI_ISL_706014\|20 | ···················· | ···················· | ····················· | ···················· | ···················· | ···················· |
| W | 2693 | hCoV-19/England/MILK-B3A2C0/2020\|EPI_ISL_706015\|20 | ···················· | ···················· | ····················· | ···················· | ···················· | ···················· |
| W | 2694 | hCoV-19/England/MILK-B3A318/2020\|EPI_ISL_706016\|20 | ···················· | ···················· | ····················· | ···················· | ···················· | ···················· |
| W | 2695 | hCoV-19/England/MILK-B3A327/2020\|EPI_ISL_706017\|20 | ···················· | ···················· | ····················· | ···················· | ···················· | ···················· |
| W | 2696 | hCoV-19/England/MILK-B3A336/2020\|EPI_ISL_706018\|20 | ···················· | ···················· | ····················· | ···················· | ······T············· | ···················· |
| W | 2697 | hCoV-19/England/MILK-B3A354/2020\|EPI_ISL_706019\|20 | ···················· | ···················· | ····················· | ···················· | ···················· | ···················· |
| W | 2698 | hCoV-19/England/MILK-B3A363/2020\|EPI_ISL_706020\|20 | ···················· | ···················· | ····················· | ···················· | ···················· | ···················· |
| W | 2699 | hCoV-19/England/MILK-B3A372/2020\|EPI_ISL_706021\|20 | ···················· | ···················· | ····················· | ···················· | ···················· | ···················· |
| W | 2700 | hCoV-19/England/MILK-B3A3AF/2020\|EPI_ISL_706022\|20 | ···················· | ···················· | ····················· | ···················· | ···················· | ···················· |
| W | 2701 | hCoV-19/England/MILK-B3A3CD/2020\|EPI_ISL_706023\|20 | ···················· | ···················· | ····················· | ···················· | ···················· | ···················· |
| W | 2702 | hCoV-19/England/MILK-B3A3EB/2020\|EPI_ISL_706024\|20 | ···················· | ···················· | ····················· | ···················· | ···················· | ···················· |
| W | 2703 | hCoV-19/England/MILK-B3A679/2020\|EPI_ISL_706025\|20 | ···················· | ···················· | ····················· | ···················· | ······T············· | ···················· |
| W | 2704 | hCoV-19/England/MILK-B3A6B5/2020\|EPI_ISL_706026\|20 | ···················· | ···················· | ····················· | ···················· | ···················· | ···················· |
| W | 2705 | hCoV-19/England/MILK-B3A6C4/2020\|EPI_ISL_706027\|20 | ···················· | ···················· | ····················· | ···················· | ···················· | ···················· |
| W | 2706 | hCoV-19/England/MILK-B3A70D/2020\|EPI_ISL_706028\|20 | ···················· | ·A·················· | ····················· | ···················· | ···················· | ···A················ |
| W | 2707 | hCoV-19/England/MILK-B3A71C/2020\|EPI_ISL_706029\|20 | ···················· | ···················· | ····················· | ···················· | ···················· | ···················· |
| W | 2708 | hCoV-19/England/MILK-B3A72B/2020\|EPI_ISL_706030\|20 | ···················· | ···················· | ····················· | ···················· | ···················· | ···················· |
| W | 2709 | hCoV-19/England/MILK-B3A749/2020\|EPI_ISL_706031\|20 | ···················· | ···················· | ····················· | ···················· | ···················· | ···················· |
| W | 2710 | hCoV-19/England/MILK-B3A758/2020\|EPI_ISL_706032\|20 | ···················· | ···················· | ····················· | ···················· | ···················· | ···················· |
| W | 2711 | hCoV-19/England/MILK-B3A767/2020\|EPI_ISL_706033\|20 | ···················· | ···················· | ····················· | ···················· | ···················· | ···················· |
| W | 2712 | hCoV-19/England/MILK-B3A776/2020\|EPI_ISL_706034\|20 | ···················· | ···················· | ····················· | ···················· | ···················· | ···················· |
| W | 2713 | hCoV-19/England/MILK-B3A794/2020\|EPI_ISL_706035\|20 | ···················· | ···················· | ····················· | ···················· | ···················· | ···················· |
| W | 2714 | hCoV-19/England/MILK-B3A7B2/2020\|EPI_ISL_706036\|20 | ···················· | ···················· | ····················· | ···················· | ···················· | ···················· |
| W | 2715 | hCoV-19/England/MILK-B3A7C1/2020\|EPI_ISL_706037\|20 | ···················· | ···················· | ····················· | ···················· | ···················· | ···················· |
| W | 2716 | hCoV-19/England/MILK-B3A7FE/2020\|EPI_ISL_706038\|20 | ···················· | ···················· | ····················· | ···················· | ···················· | ···················· |
| W | 2717 | hCoV-19/England/MILK-B3A819/2020\|EPI_ISL_706039\|20 | ···················· | ···················· | ····················· | ···················· | ···················· | ···················· |
| W | 2718 | hCoV-19/England/MILK-B3A837/2020\|EPI_ISL_706040\|20 | ···················· | ···················· | ····················· | ···················· | ···················· | ···················· |
| W | 2719 | hCoV-19/England/MILK-B3AA5F/2020\|EPI_ISL_706041\|20 | ···················· | ···················· | ····················· | ···················· | ···················· | ···················· |
| W | 2720 | hCoV-19/England/MILK-B3AA6E/2020\|EPI_ISL_706042\|20 | ···················· | ···················· | ····················· | ···················· | ···················· | ···················· |
| W | 2721 | hCoV-19/England/MILK-B3AA9B/2020\|EPI_ISL_706043\|20 | ···················· | ···················· | ····················· | ···················· | ······T············· | ···················· |
| W | 2722 | hCoV-19/England/MILK-B3AAC8/2020\|EPI_ISL_706044\|20 | ···················· | ···················· | ····················· | ···················· | ···················· | ···················· |
| W | 2723 | hCoV-19/England/MILK-B3AAD7/2020\|EPI_ISL_706045\|20 | ···················· | ···················· | ····················· | ···················· | ···················· | ···················· |
| W | 2724 | hCoV-19/England/MILK-B3AAF5/2020\|EPI_ISL_706046\|20 | ···················· | ···················· | ····················· | ···················· | ······T············· | ···················· |
| W | 2725 | hCoV-19/England/MILK-B3AB01/2020\|EPI_ISL_706047\|20 | ···················· | ···················· | ····················· | ···················· | ······T············· | ···················· |
| W | 2726 | hCoV-19/England/MILK-B3AB10/2020\|EPI_ISL_706048\|20 | ···················· | ···················· | ····················· | ···················· | ···················· | ···················· |
| W | 2727 | hCoV-19/England/MILK-B3B14A/2020\|EPI_ISL_706049\|20 | ···················· | ···················· | ····················· | ···················· | ···················· | ···················· |
| W | 2728 | hCoV-19/England/MILK-B3B159/2020\|EPI_ISL_706050\|20 | ···················· | ·A·················· | ····················· | ···················· | ···················· | ···A················ |
| W | 2729 | hCoV-19/England/MILK-B3B168/2020\|EPI_ISL_706051\|20 | ···················· | ···················· | ····················· | ···················· | ···················· | ···················· |
| W | 2730 | hCoV-19/England/MILK-B3B177/2020\|EPI_ISL_706052\|20 | ···················· | ···················· | ····················· | ···················· | ···················· | ···················· |

| | | | | | | | | |
|---|---|---|---|---|---|---|---|---|
| W | 2731 | hCoV-19/England/MILK-B3B1C2/2020\|EPI_ISL_706053\|20 | ···················· | ···················· | ···················· | ···················· | ···················· | ···················· |
| W | 2732 | hCoV-19/England/MILK-B3B20B/2020\|EPI_ISL_706054\|20 | ···················· | ···················· | ···················· | ···················· | ···················· | ···················· |
| W | 2733 | hCoV-19/England/MILK-B3B238/2020\|EPI_ISL_706055\|20 | ···················· | ···················· | ···················· | ···················· | ···················· | ···················· |
| W | 2734 | hCoV-19/England/MILK-B3B247/2020\|EPI_ISL_706056\|20 | ···················· | ···················· | ···················· | ···················· | ···················· | ···················· |
| W | 2735 | hCoV-19/England/MILK-B3B265/2020\|EPI_ISL_706057\|20 | ···················· | ···················· | ···················· | ···················· | ···················· | ···A················ |
| W | 2736 | hCoV-19/England/MILK-B3B353/2020\|EPI_ISL_706058\|20 | ···················· | ···················· | ···················· | ···················· | ···················· | ···················· |
| W | 2737 | hCoV-19/England/MILK-B3B380/2020\|EPI_ISL_706059\|20 | ···················· | ···················· | ···················· | ···················· | ···················· | ···················· |
| W | 2738 | hCoV-19/England/MILK-B3B3F9/2020\|EPI_ISL_706060\|20 | ···················· | ···················· | ···················· | ···················· | ···················· | ···················· |
| W | 2739 | hCoV-19/England/MILK-B3B4AB/2020\|EPI_ISL_706061\|20 | ···················· | ···················· | ···················· | ···················· | ···················· | ···················· |
| W | 2740 | hCoV-19/England/MILK-B3B4C9/2020\|EPI_ISL_706062\|20 | ···················· | ···················· | ···················· | ···················· | ···················· | ···················· |
| W | 2741 | hCoV-19/England/MILK-B3B55D/2020\|EPI_ISL_706063\|20 | ···················· | ···················· | ···················· | ···················· | ···················· | ···················· |
| W | 2742 | hCoV-19/England/MILK-B3B57B/2020\|EPI_ISL_706064\|20 | ···················· | ···················· | ···················· | ···················· | ···················· | ···················· |
| W | 2743 | hCoV-19/England/MILK-B3B599/2020\|EPI_ISL_706065\|20 | ···················· | ···················· | ···················· | ···················· | ···················· | ···················· |
| W | 2744 | hCoV-19/England/MILK-B3B5B7/2020\|EPI_ISL_706066\|20 | ···················· | ···················· | ···················· | ···················· | ···················· | ···················· |
| W | 2745 | hCoV-19/England/MILK-B3B5C6/2020\|EPI_ISL_706067\|20 | ···················· | ···················· | ···················· | ···················· | ···················· | ···················· |
| W | 2746 | hCoV-19/England/MILK-B3B5E4/2020\|EPI_ISL_706068\|20 | ···················· | ···················· | ···················· | ···················· | ···················· | ···················· |
| W | 2747 | hCoV-19/England/MILK-B3B60F/2020\|EPI_ISL_706069\|20 | ···················· | ···················· | ···················· | ···················· | ···················· | ···················· |
| W | 2748 | hCoV-19/England/MILK-B3B61E/2020\|EPI_ISL_706070\|20 | ···················· | ···················· | ···················· | ···················· | ···················· | ···················· |
| W | 2749 | hCoV-19/England/MILK-B3B62D/2020\|EPI_ISL_706071\|20 | ···················· | ···················· | ···················· | ···················· | ···················· | ···················· |
| W | 2750 | hCoV-19/England/MILK-B3B64B/2020\|EPI_ISL_706072\|20 | ···················· | ·A·················· | ···················· | ···················· | ···················· | ···A················ |
| W | 2751 | hCoV-19/England/MILK-B3B65A/2020\|EPI_ISL_706073\|20 | ···················· | ···················· | ···················· | ···················· | ······T············· | ···················· |
| W | 2752 | hCoV-19/England/MILK-B3B678/2020\|EPI_ISL_706074\|20 | ···················· | ···················· | ···················· | ···················· | ···················· | ···················· |
| W | 2753 | hCoV-19/England/MILK-B3B687/2020\|EPI_ISL_706075\|20 | ···················· | ···················· | ···················· | ···················· | ···················· | ···················· |
| W | 2754 | hCoV-19/England/MILK-B3B696/2020\|EPI_ISL_706076\|20 | ···················· | ···················· | ···················· | ···················· | ···················· | ···················· |
| W | 2755 | hCoV-19/England/MILK-B3B6B4/2020\|EPI_ISL_706077\|20 | ···················· | ···················· | ···················· | ···················· | ···················· | ···················· |
| W | 2756 | hCoV-19/England/MILK-B3B6C3/2020\|EPI_ISL_706078\|20 | ···················· | ···················· | ···················· | ···················· | ···················· | ···················· |
| W | 2757 | hCoV-19/England/MILK-B3B6E1/2020\|EPI_ISL_706079\|20 | ···················· | ···················· | ···················· | ···················· | ···················· | ···················· |
| W | 2758 | hCoV-19/England/MILK-B3B70C/2020\|EPI_ISL_706080\|20 | ···················· | ···················· | ···················· | ···················· | ···················· | ···················· |
| W | 2759 | hCoV-19/England/MILK-B3B881/2020\|EPI_ISL_706081\|20 | ···················· | ···················· | ···················· | ···················· | ···················· | ···················· |
| W | 2760 | hCoV-19/England/MILK-B3B890/2020\|EPI_ISL_706082\|20 | ···················· | ···················· | ···················· | ···················· | ···················· | ···················· |
| W | 2761 | hCoV-19/England/MILK-B3B8AF/2020\|EPI_ISL_706083\|20 | ···················· | ···················· | ···················· | ···················· | ···················· | ···················· |
| W | 2762 | hCoV-19/England/MILK-B3B8BE/2020\|EPI_ISL_706084\|20 | ···················· | ···················· | ···················· | ···················· | ···················· | ···················· |
| W | 2763 | hCoV-19/England/MILK-B3B906/2020\|EPI_ISL_706085\|20 | ···················· | ···················· | ···················· | ···················· | ···················· | ···················· |
| W | 2764 | hCoV-19/England/MILK-B3B915/2020\|EPI_ISL_706086\|20 | ···················· | ···················· | ···················· | ···················· | ···················· | ···················· |
| W | 2765 | hCoV-19/England/MILK-B3B924/2020\|EPI_ISL_706087\|20 | ···················· | ···················· | ···················· | ···················· | ···················· | ···················· |
| W | 2766 | hCoV-19/England/MILK-B3B933/2020\|EPI_ISL_706088\|20 | ···················· | ···················· | ···················· | ···················· | ···················· | ···················· |
| W | 2767 | hCoV-19/England/MILK-B3B951/2020\|EPI_ISL_706089\|20 | ···················· | ···················· | ···················· | ···················· | ···················· | ···················· |
| W | 2768 | hCoV-19/England/MILK-B3B98E/2020\|EPI_ISL_706090\|20 | ···················· | ···················· | ···················· | ···················· | ···················· | ···················· |
| W | 2769 | hCoV-19/England/MILK-B3B9CA/2020\|EPI_ISL_706091\|20 | ···················· | ···················· | ···················· | ···················· | ···················· | ···················· |
| W | 2770 | hCoV-19/England/MILK-B3B9F7/2020\|EPI_ISL_706092\|20 | ···················· | ···················· | ···················· | ···················· | ···················· | ···················· |
| W | 2771 | hCoV-19/England/MILK-B3BA21/2020\|EPI_ISL_706093\|20 | ···················· | ···················· | ···················· | ···················· | ···················· | ···················· |
| W | 2772 | hCoV-19/England/MILK-B3BAF4/2020\|EPI_ISL_706094\|20 | ···················· | ···················· | ···················· | ···················· | ···················· | ···················· |
| W | 2773 | hCoV-19/England/MILK-B3BB1F/2020\|EPI_ISL_706095\|20 | ···················· | ···················· | ···················· | ···················· | ···················· | ···················· |
| W | 2774 | hCoV-19/England/MILK-B3BB2E/2020\|EPI_ISL_706096\|20 | ···················· | ···················· | ···················· | ···················· | ···················· | ···················· |
| W | 2775 | hCoV-19/England/MILK-B3BB3D/2020\|EPI_ISL_706097\|20 | ···················· | ···················· | ···················· | ···················· | ···················· | ···················· |
| W | 2776 | hCoV-19/England/MILK-B3BB6A/2020\|EPI_ISL_706098\|20 | ···················· | ···················· | ···················· | ···················· | ······T············· | ···················· |
| W | 2777 | hCoV-19/England/MILK-B3BE25/2020\|EPI_ISL_706099\|20 | ···················· | ···················· | ···················· | ···················· | ···················· | ···················· |
| W | 2778 | hCoV-19/USA/TX-DSHS-1685/2020\|EPI_ISL_708646\|2020- | ···················· | ···················· | ···················· | ···················· | ···················· | ···················· |
| W | 2779 | hCoV-19/USA/TX-DSHS-1687/2020\|EPI_ISL_708647\|2020- | ···················· | ···················· | ···················· | ···················· | ···················· | ···················· |
| W | 2780 | hCoV-19/USA/TX-DSHS-1698/2020\|EPI_ISL_708648\|2020- | ···················· | ···················· | ···················· | ···················· | ···················· | ···················· |
| W | 2781 | hCoV-19/USA/TX-DSHS-1723/2020\|EPI_ISL_708649\|2020- | ···················· | ···················· | ···················· | ···················· | ···················· | ···················· |
| W | 2782 | hCoV-19/Gibraltar/205000624/2020\|EPI_ISL_709960\|20 | ···················· | ···················· | ···················· | ···················· | ···················· | ···················· |
| W | 2783 | hCoV-19/Gibraltar/205000625/2020\|EPI_ISL_709961\|20 | ···················· | ···················· | ···················· | ···················· | ···················· | ···················· |
| W | 2784 | hCoV-19/Gibraltar/205000626/2020\|EPI_ISL_709962\|20 | ···················· | ···················· | ···················· | ···················· | ···················· | ···················· |
| W | 2785 | hCoV-19/Gibraltar/205000627/2020\|EPI_ISL_709963\|20 | ···················· | ···················· | ···················· | ···················· | ···················· | ···················· |
| W | 2786 | hCoV-19/Gibraltar/205000628/2020\|EPI_ISL_709964\|20 | ···················· | ···················· | ···················· | ···················· | ···················· | ···················· |
| W | 2787 | hCoV-19/Gibraltar/205000630/2020\|EPI_ISL_709965\|20 | ···················· | ···················· | ···················· | ···················· | ···················· | ···················· |
| W | 2788 | hCoV-19/Gibraltar/205000631/2020\|EPI_ISL_709966\|20 | ···················· | ···················· | ···················· | ···················· | ···················· | ···················· |
| W | 2789 | hCoV-19/Gibraltar/205000632/2020\|EPI_ISL_709967\|20 | ···················· | ···················· | ···················· | ···················· | ···················· | ···················· |
| W | 2790 | hCoV-19/Gibraltar/205000633/2020\|EPI_ISL_709968\|20 | ···················· | ···················· | ···················· | ···················· | ···················· | ···················· |
| W | 2791 | hCoV-19/Gibraltar/205000634/2020\|EPI_ISL_709969\|20 | ···················· | ···················· | ···················· | ···················· | ···················· | ···················· |
| W | 2792 | hCoV-19/Gibraltar/205000635/2020\|EPI_ISL_709970\|20 | ···················· | ···················· | ···················· | ···················· | ···················· | ···················· |
| W | 2793 | hCoV-19/Gibraltar/205000636/2020\|EPI_ISL_709971\|20 | ···················· | ···················· | ···················· | ···················· | ···················· | ···················· |
| W | 2794 | hCoV-19/Gibraltar/205000637/2020\|EPI_ISL_709972\|20 | ···················· | ···················· | ···················· | ···················· | ···················· | ···················· |
| W | 2795 | hCoV-19/Gibraltar/205000638/2020\|EPI_ISL_709973\|20 | ···················· | ···················· | ···················· | ···················· | ···················· | ···················· |
| W | 2796 | hCoV-19/Gibraltar/205000639/2020\|EPI_ISL_709974\|20 | ···················· | ···················· | ···················· | ···················· | ···················· | ···················· |
| W | 2797 | hCoV-19/Gibraltar/205000640/2020\|EPI_ISL_709975\|20 | ···················· | ···················· | ···················· | ···················· | ···················· | ···················· |
| W | 2798 | hCoV-19/Gibraltar/205000641/2020\|EPI_ISL_709976\|20 | ···················· | ···················· | ···················· | ···················· | ···················· | ···················· |
| W | 2799 | hCoV-19/Gibraltar/205000642/2020\|EPI_ISL_709977\|20 | ···················· | ···················· | ···················· | ···················· | ···················· | ···················· |
| W | 2800 | hCoV-19/Gibraltar/205000643/2020\|EPI_ISL_709978\|20 | ···················· | ···················· | ···················· | ···················· | ···················· | ···················· |
| W | 2801 | hCoV-19/Gibraltar/205000644/2020\|EPI_ISL_709979\|20 | ···················· | ···················· | ···················· | ···················· | ···················· | ···················· |
| W | 2802 | hCoV-19/Gibraltar/205000646/2020\|EPI_ISL_709981\|20 | ···················· | ···················· | ···················· | ···················· | ···················· | ···················· |
| W | 2803 | hCoV-19/Gibraltar/205000647/2020\|EPI_ISL_709982\|20 | ···················· | ···················· | ···················· | ···················· | ···················· | ···············A···· |
| W | 2804 | hCoV-19/Gibraltar/205000648/2020\|EPI_ISL_709983\|20 | ···················· | ···················· | ···················· | ···················· | ···················· | ···················· |
| W | 2805 | hCoV-19/Gibraltar/205000649/2020\|EPI_ISL_709984\|20 | ···················· | ···················· | ···················· | ···················· | ···················· | ···················· |
| W | 2806 | hCoV-19/Gibraltar/205000658/2020\|EPI_ISL_709989\|20 | ···················· | ···················· | ···················· | ···················· | ···················· | ···················· |
| W | 2807 | hCoV-19/Gibraltar/205000659/2020\|EPI_ISL_709990\|20 | ···················· | ···················· | ···················· | ···················· | ···················· | ···················· |
| W | 2808 | hCoV-19/Gibraltar/205000660/2020\|EPI_ISL_709991\|20 | ···················· | ···················· | ···················· | ···················· | ···················· | ···················· |
| W | 2809 | hCoV-19/Gibraltar/205000661/2020\|EPI_ISL_709992\|20 | ···················· | ···················· | ···················· | ···················· | ···················· | ···················· |
| W | 2810 | hCoV-19/Gibraltar/205000666/2020\|EPI_ISL_709993\|20 | ···················· | ···················· | ···················· | ···················· | ···················· | ···················· |

```
W    2811 hCoV-19/Gibraltar/205000667/2020|EPI_ISL_709994|20    ····················  ····················  ·····················  ····················  ····················  ····················
W    2812 hCoV-19/Gibraltar/205000669/2020|EPI_ISL_709995|20    ····················  ····················  ·····················  ····················  ····················  ····················
W    2813 hCoV-19/Gibraltar/205000670/2020|EPI_ISL_709996|20    ····················  ····················  ·····················  ····················  ····················  ····················
W    2814 hCoV-19/Gibraltar/205000671/2020|EPI_ISL_709997|20    ····················  ····················  ·····················  ····················  ····················  ····················
W    2815 hCoV-19/Gibraltar/205000672/2020|EPI_ISL_709998|20    ····················  ····················  ·····················  ····················  ····················  ····················
W    2816 hCoV-19/Gibraltar/205000673/2020|EPI_ISL_709999|20    ····················  ····················  ·····················  ····················  ····················  ····················
W    2817 hCoV-19/USA/CA-LACPHL-AB00001/2020|EPI_ISL_694046|    ····················  ····················  ·····················  ····················  ····················  ····················
W    2818 hCoV-19/USA/AZ-TG266871/2020|EPI_ISL_694047|2020-0    ····················  ····················  ·····················  ····················  ····················  ····················
W    2819 hCoV-19/USA/AZ-TG269787/2020|EPI_ISL_694048|2020-0    ····················  ····················  ·····················  ····················  ····················  ····················
W    2820 hCoV-19/USA/AZ-TG269891/2020|EPI_ISL_694049|2020-0    ····················  ····A···············  ·····················  ····················  ····················  ····················
W    2821 hCoV-19/USA/AZ-TG270095/2020|EPI_ISL_694050|2020-0    ····················  ····················  ·····················  ····················  ····················  ····················
W    2822 hCoV-19/USA/AZ-TG271575/2020|EPI_ISL_694052|2020-0    ····················  ····················  ·····················  ····················  ····················  ····················
W    2823 hCoV-19/USA/AZ-TG273320/2020|EPI_ISL_694053|2020-0    ····················  ····················  ·····················  ····················  ····················  ····················
W    2824 hCoV-19/USA/AZ-TG273328/2020|EPI_ISL_694054|2020-0    ····················  ····················  ·····················  ····················  ····················  ····················
W    2825 hCoV-19/USA/AZ-TG273340/2020|EPI_ISL_694055|2020-0    ····················  ····················  ·····················  ····················  ····················  ····················
W    2826 hCoV-19/USA/AZ-TG273342/2020|EPI_ISL_694056|2020-0    ····················  ····················  ·····················  ····················  ····················  ····················
W    2827 hCoV-19/USA/AZ-TG273344/2020|EPI_ISL_694057|2020-0    ····················  ····················  ·····················  ····················  ····················  ····················
W    2828 hCoV-19/USA/AZ-TG273354/2020|EPI_ISL_694058|2020-0    ····················  ····················  ·····················  ····················  ····················  ····················
W    2829 hCoV-19/USA/AZ-TG273356/2020|EPI_ISL_694059|2020-0    ····················  ····················  ·····················  ····················  ····················  ····················
W    2830 hCoV-19/USA/AZ-TG274698/2020|EPI_ISL_694060|2020-0    ····················  ····················  ·····················  ····················  ····················  ····················
W    2831 hCoV-19/USA/AZ-TG274709/2020|EPI_ISL_694061|2020-0    ····················  ····················  ·····················  ····················  ····················  ····················
W    2832 hCoV-19/USA/AZ-TG274711/2020|EPI_ISL_694062|2020-0    ····················  ····················  ·····················  ····················  ····················  ····················
W    2833 hCoV-19/USA/AZ-TG274723/2020|EPI_ISL_694064|2020-0    ····················  ····················  ·····················  ····················  ····················  ····················
W    2834 hCoV-19/USA/AZ-TG274744/2020|EPI_ISL_694066|2020-0    ····················  ····················  ·····················  ····················  ····················  ····················
W    2835 hCoV-19/USA/AZ-TG274746/2020|EPI_ISL_694067|2020-0    ····················  ····················  ·····················  ····················  ····················  ····················
W    2836 hCoV-19/USA/AZ-TG274748/2020|EPI_ISL_694068|2020-0    ····················  ····················  ·····················  ····················  ····T···············  ····················
W    2837 hCoV-19/USA/AZ-TG274750/2020|EPI_ISL_694069|2020-0    ····················  ····················  ·····················  ····················  ····················  ····················
W    2838 hCoV-19/USA/AZ-TG274754/2020|EPI_ISL_694070|2020-0    ····················  ····················  ·····················  ····················  ····················  ····················
W    2839 hCoV-19/USA/AZ-TG274756/2020|EPI_ISL_694071|2020-0    ····················  ····················  ·····················  ····················  ····················  ····················
W    2840 hCoV-19/USA/AZ-TG274763/2020|EPI_ISL_694072|2020-0    ····················  ····················  ·····················  ····················  ····················  ····················
W    2841 hCoV-19/USA/AZ-TG274765/2020|EPI_ISL_694073|2020-0    ····················  ····················  ·····················  ····················  ····················  ····················
W    2842 hCoV-19/USA/AZ-TG274767/2020|EPI_ISL_694074|2020-0    ····················  ····················  ·····················  ····················  ····················  ····················
W    2843 hCoV-19/USA/AZ-TG274769/2020|EPI_ISL_694075|2020-0    ····················  ····················  ·····················  ····················  ····················  ····················
W    2844 hCoV-19/USA/AZ-TG274779/2020|EPI_ISL_694077|2020-0    ····················  ····················  ·····················  ····················  ····················  ····················
W    2845 hCoV-19/USA/AZ-TG274786/2020|EPI_ISL_694078|2020-0    ····················  ····················  ·····················  ····················  ····················  ····················
W    2846 hCoV-19/USA/AZ-TG274788/2020|EPI_ISL_694079|2020-0    ····················  ····················  ·····················  ····················  ····················  ····················
W    2847 hCoV-19/USA/AZ-TG274800/2020|EPI_ISL_694082|2020-0    ····················  ····················  ·····················  ····················  ····················  ····················
W    2848 hCoV-19/USA/AZ-TG274810/2020|EPI_ISL_694083|2020-0    ····················  ····················  ·····················  ····················  ····················  ····················
W    2849 hCoV-19/USA/AZ-TG276124/2020|EPI_ISL_694084|2020-0    ····················  ····················  ·····················  ····················  ····················  ····················
W    2850 hCoV-19/USA/AZ-TG276128/2020|EPI_ISL_694085|2020-0    ····················  ····················  ·····················  ····················  ····················  ····················
W    2851 hCoV-19/USA/AZ-TG276130/2020|EPI_ISL_694086|2020-0    ····················  ·····A··············  ·····················  ····················  ····················  ····················
W    2852 hCoV-19/USA/AZ-TG276132/2020|EPI_ISL_694087|2020-0    ····················  ····················  ·····················  ····················  ····················  ····················
W    2853 hCoV-19/USA/AZ-TG276136/2020|EPI_ISL_694088|2020-0    ····················  ····················  ·····················  ····················  ····················  ····················
W    2854 hCoV-19/USA/AZ-TG276138/2020|EPI_ISL_694089|2020-0    ····················  ····················  ·····················  ····················  ····················  ····················
W    2855 hCoV-19/USA/AZ-TG276142/2020|EPI_ISL_694090|2020-0    ····················  ····················  ·····················  ····················  ····················  ····················
W    2856 hCoV-19/USA/AZ-TG276152/2020|EPI_ISL_694093|2020-0    ····················  ····················  ·····················  ····················  ····················  ····················
W    2857 hCoV-19/USA/AZ-TG276158/2020|EPI_ISL_694094|2020-0    ····················  ····················  ·····················  ····················  ····················  ····················
W    2858 hCoV-19/USA/AZ-TG276171/2020|EPI_ISL_694095|2020-0    ····················  ····················  ·····················  ····················  ····················  ····················
W    2859 hCoV-19/USA/AZ-TG276173/2020|EPI_ISL_694096|2020-0    ····················  ····················  ·····················  ····················  ····················  ····················
W    2860 hCoV-19/USA/AZ-TG276177/2020|EPI_ISL_694097|2020-0    ····················  ····················  ·····················  ····················  ····················  ····················
W    2861 hCoV-19/USA/AZ-TG276179/2020|EPI_ISL_694098|2020-0    ····················  ····················  ·····················  ····················  ····················  ····················
W    2862 hCoV-19/USA/AZ-TG276181/2020|EPI_ISL_694099|2020-0    ····················  ····················  ·····················  ····················  ····················  ····················
W    2863 hCoV-19/USA/AZ-TG508113/2020|EPI_ISL_695301|2020-0    ····················  ·A··················  ·····················  ····················  ····················  ····················
W    2864 hCoV-19/USA/AZ-TG508838/2020|EPI_ISL_695303|2020-0    ····················  ····················  ·····················  ····················  ····················  ····················
W    2865 hCoV-19/USA/AZ-TG513389/2020|EPI_ISL_695308|2020-0    ····················  ·A··················  ·····················  ····················  ····················  ····················
W    2866 hCoV-19/USA/AZ-TG513833/2020|EPI_ISL_695311|2020-0    ····················  ····················  ·····················  ····················  ····················  ····················
W    2867 hCoV-19/USA/AZ-TG516126/2020|EPI_ISL_695312|2020-0    ····················  ····················  ·····················  ····················  ····················  ····················
W    2868 hCoV-19/USA/AZ-TG516154/2020|EPI_ISL_695313|2020-0    ····················  ····················  ·····················  ····················  ····················  ····················
W    2869 hCoV-19/USA/AZ-TG517469/2020|EPI_ISL_695328|2020-0    ····················  ·A··················  ·····················  ····················  ····················  ····················
W    2870 hCoV-19/USA/AZ-TG517520/2020|EPI_ISL_695345|2020-0    ····················  ·A··················  ·····················  ····················  ····················  ····················
W    2871 hCoV-19/USA/AZ-TG519128/2020|EPI_ISL_695351|2020-0    ····················  ····················  ·····················  ····················  ····················  ····················
W    2872 hCoV-19/USA/AZ-TG519139/2020|EPI_ISL_695354|2020-0    ····················  ····················  ·····················  ····················  ····················  ····················
W    2873 hCoV-19/USA/AZ-TG519143/2020|EPI_ISL_695355|2020-0    ····················  ····················  ·····················  ····················  ····················  ····················
W    2874 hCoV-19/USA/AZ-TG519162/2020|EPI_ISL_695360|2020-0    ····················  ·············A······  ·····················  ····················  ····················  ····················
W    2875 hCoV-19/USA/AZ-TG519163/2020|EPI_ISL_695361|2020-0    ····················  ····················  ·····················  ····················  ····················  ····················
W    2876 hCoV-19/USA/AZ-TG519164/2020|EPI_ISL_695362|2020-0    ····················  ······N·············  ·····················  ····················  ····················  ····················
W    2877 hCoV-19/USA/AZ-TG519187/2020|EPI_ISL_695367|2020-0    ····················  ·A··················  ·····················  ····················  ····················  ····················
W    2878 hCoV-19/USA/AZ-TG519189/2020|EPI_ISL_695368|2020-0    ····················  ·A··················  ·····················  ····················  ····················  ····················
W    2879 hCoV-19/USA/AZ-TG519200/2020|EPI_ISL_695370|2020-0    ····················  ·A··················  ·····················  ····················  ····················  ····················
W    2880 hCoV-19/USA/AZ-TG521369/2020|EPI_ISL_695376|2020-0    ····················  ····················  ·····················  ····················  ····················  ····················
W    2881 hCoV-19/USA/AZ-TG521376/2020|EPI_ISL_695378|2020-0    ····················  ·A··················  ·····················  ····················  ····················  ····················
W    2882 hCoV-19/USA/AZ-TG532044/2020|EPI_ISL_695397|2020-0    ····················  ·············A······  ·····················  ····················  ····················  ····················
W    2884 hCoV-19/England/ALDP-BB9311/2020|EPI_ISL_702548|20    ····················  ····················  ·····················  ····················  ····················  ····················
W    2885 hCoV-19/England/ALDP-BBABFA/2020|EPI_ISL_702570|20    ····················  ····················  ·····················  ····················  ····················  ····················
W    2886 hCoV-19/England/ALDP-BBA5B0/2020|EPI_ISL_702572|20    ····················  ····················  ·····················  ····················  ····················  ····················
W    2887 hCoV-19/England/ALDP-BB9575/2020|EPI_ISL_702573|20    ····················  ····················  ·····················  ····················  ····················  ····················
W    2888 hCoV-19/England/ALDP-BBADF4/2020|EPI_ISL_702575|20    ····················  ·A··················  ·····················  ····················  ····················  ····················
W    2889 hCoV-19/England/ALDP-BBB24F/2020|EPI_ISL_702576|20    ····················  ····················  ·····················  ····················  ····················  ····················
W    2890 hCoV-19/England/ALDP-BBB5CE/2020|EPI_ISL_702578|20    ····················  ····················  ·····················  ····················  ····················  ····················
W    2891 hCoV-19/England/ALDP-BBB3E2/2020|EPI_ISL_702580|20    ····················  ····················  ·····················  ····················  ····················  ····················
```

| | | | | | | | |
|---|---|---|---|---|---|---|---|
| W | 2892 hCoV-19/England/ALDP-BBABCD/2020\|EPI_ISL_702581\|20 | ···················· | ···················· | ····················· | ···················· | ···················· | ···················· |
| W | 2893 hCoV-19/England/ALDP-BB9663/2020\|EPI_ISL_702583\|20 | ···················· | ···················· | ····················· | ···················· | ···················· | ···················· |
| W | 2894 hCoV-19/England/ALDP-BB94E1/2020\|EPI_ISL_702588\|20 | ···················· | ···················· | ····················· | ···················· | ···················· | ···················· |
| W | 2895 hCoV-19/England/ALDP-BBA6AE/2020\|EPI_ISL_702589\|20 | ···················· | ·A·················· | ····················· | ···················· | ···················· | ···A················ |
| W | 2896 hCoV-19/England/ALDP-BBA93C/2020\|EPI_ISL_702592\|20 | ···················· | ···················· | ····················· | ···················· | ···················· | ···················· |
| W | 2897 hCoV-19/England/ALDP-BB4932/2020\|EPI_ISL_702593\|20 | ···················· | ···················· | ····················· | ···················· | ······T············· | ···················· |
| W | 2898 hCoV-19/England/ALDP-BBB41C/2020\|EPI_ISL_702595\|20 | ···················· | ···················· | ····················· | ···················· | ···················· | ···················· |
| W | 2899 hCoV-19/England/ALDP-BB4E7F/2020\|EPI_ISL_702596\|20 | ···················· | ···················· | ····················· | ···················· | ···················· | ···················· |
| W | 2900 hCoV-19/England/ALDP-BB9566/2020\|EPI_ISL_702598\|20 | ···················· | ···················· | ····················· | ···················· | ···················· | ···················· |
| W | 2901 hCoV-19/England/ALDP-BB929C/2020\|EPI_ISL_702599\|20 | ···················· | ···················· | ····················· | ···················· | ···················· | ···················· |
| W | 2902 hCoV-19/England/ALDP-BB9C07/2020\|EPI_ISL_702601\|20 | ···················· | ···················· | ····················· | ···················· | ······T············· | ···················· |
| W | 2903 hCoV-19/England/ALDP-BBA5ED/2020\|EPI_ISL_702603\|20 | ···················· | ···················· | ····················· | ···················· | ···················· | ···················· |
| W | 2904 hCoV-19/England/ALDP-BB4EAC/2020\|EPI_ISL_702604\|20 | ···················· | ·A·················· | ····················· | ···················· | ···················· | ···················· |
| W | 2905 hCoV-19/England/ALDP-BB9C43/2020\|EPI_ISL_702606\|20 | ···················· | ···················· | ····················· | ···················· | ···················· | ···················· |
| W | 2906 hCoV-19/England/ALDP-BB4EBB/2020\|EPI_ISL_702608\|20 | ···················· | ···················· | ····················· | ···················· | ···················· | ···················· |
| W | 2907 hCoV-19/England/ALDP-BB4914/2020\|EPI_ISL_702609\|20 | ···················· | ···················· | ····················· | ···················· | ···················· | ···················· |
| W | 2908 hCoV-19/England/ALDP-BB4DCD/2020\|EPI_ISL_702611\|20 | ···················· | ···················· | ····················· | ···················· | ···················· | ···················· |
| W | 2909 hCoV-19/England/ALDP-BB5719/2020\|EPI_ISL_702627\|20 | ···················· | ···················· | ····················· | ···················· | ···················· | ···················· |
| W | 2910 hCoV-19/England/ALDP-BB3432/2020\|EPI_ISL_702629\|20 | ···················· | ···················· | ····················· | ···················· | ···················· | ···················· |
| W | 2911 hCoV-19/England/ALDP-BBAD30/2020\|EPI_ISL_702630\|20 | ···················· | ···················· | ····················· | ···················· | ···················· | ···················· |
| W | 2912 hCoV-19/England/ALDP-BB99A6/2020\|EPI_ISL_702632\|20 | ···················· | ···················· | ····················· | ···················· | ···················· | ···················· |
| W | 2913 hCoV-19/England/ALDP-BB362D/2020\|EPI_ISL_702634\|20 | ···················· | ···················· | ····················· | ···················· | ···················· | ···················· |
| W | 2914 hCoV-19/England/ALDP-BB356C/2020\|EPI_ISL_702635\|20 | ···················· | ···················· | ····················· | ···················· | ···················· | ···················· |
| W | 2915 hCoV-19/England/ALDP-BB3195/2020\|EPI_ISL_702637\|20 | ···················· | ···················· | ····················· | ···················· | ···················· | ···················· |
| W | 2916 hCoV-19/Wales/ALDP-BB9469/2020\|EPI_ISL_702641\|2020 | ···················· | ···················· | ····················· | ···················· | ···················· | ···················· |
| W | 2917 hCoV-19/England/ALDP-BB27D0/2020\|EPI_ISL_702643\|20 | ···················· | ···················· | ····················· | ···················· | ···················· | ···················· |
| W | 2918 hCoV-19/England/ALDP-BB8C17/2020\|EPI_ISL_702645\|20 | ···················· | ···················· | ····················· | ···················· | ···················· | ···················· |
| W | 2919 hCoV-19/England/ALDP-BB9B0A/2020\|EPI_ISL_702646\|20 | ···················· | ···················· | ····················· | ···················· | ···················· | ···················· |
| W | 2920 hCoV-19/England/QEUH-BCB5F8/2020\|EPI_ISL_703877\|20 | ···················· | ···················· | ····················· | ···················· | ···················· | ···················· |
| W | 2921 hCoV-19/England/QEUH-BC3C28/2020\|EPI_ISL_703900\|20 | ···················· | ···················· | ····················· | ···················· | ···················· | ···················· |
| W | 2922 hCoV-19/England/QEUH-BC39F4/2020\|EPI_ISL_703901\|20 | ···················· | ···················· | ····················· | ···················· | ···················· | ···················· |
| W | 2923 hCoV-19/England/QEUH-BCBC8A/2020\|EPI_ISL_703904\|20 | ···················· | ···················· | ····················· | ···················· | ···················· | ···················· |
| W | 2924 hCoV-19/England/QEUH-BC37EB/2020\|EPI_ISL_703906\|20 | ···················· | ···················· | ····················· | ···················· | ···················· | ···················· |
| W | 2925 hCoV-19/England/QEUH-BCBDA5/2020\|EPI_ISL_703907\|20 | ···················· | ···················· | ····················· | ···················· | ···················· | ···················· |
| W | 2926 hCoV-19/England/QEUH-BC4BFD/2020\|EPI_ISL_703909\|20 | ···················· | ···················· | ····················· | ···················· | ···················· | ···················· |
| W | 2927 hCoV-19/England/QEUH-BCB868/2020\|EPI_ISL_703910\|20 | ···················· | ···················· | ····················· | ···················· | ···················· | ···················· |
| W | 2928 hCoV-19/England/QEUH-BCBCF3/2020\|EPI_ISL_703912\|20 | ···················· | ···················· | ····················· | ···················· | ······T············· | ···················· |
| W | 2929 hCoV-19/England/QEUH-BC4B2A/2020\|EPI_ISL_703917\|20 | ···················· | ···················· | ····················· | ···················· | ···················· | ···················· |
| W | 2930 hCoV-19/England/QEUH-BCC6F4/2020\|EPI_ISL_703919\|20 | ···················· | ·A·················· | ····················· | ···················· | ···················· | ···················· |
| W | 2931 hCoV-19/England/QEUH-BC3CCE/2020\|EPI_ISL_703925\|20 | ···················· | ···················· | ····················· | ···················· | ···················· | ···················· |
| W | 2932 hCoV-19/England/QEUH-BCC0F6/2020\|EPI_ISL_703928\|20 | ···················· | ···················· | ····················· | ···················· | ···················· | ···················· |
| W | 2933 hCoV-19/England/QEUH-BC3A1F/2020\|EPI_ISL_703930\|20 | ···················· | ···················· | ····················· | ···················· | ···················· | ···················· |
| W | 2934 hCoV-19/England/QEUH-BCB9ED/2020\|EPI_ISL_703931\|20 | ···················· | ···················· | ····················· | ···················· | ···················· | ···················· |
| W | 2935 hCoV-19/England/QEUH-BCB622/2020\|EPI_ISL_703934\|20 | ···················· | ···················· | ····················· | ···················· | ···················· | ···················· |
| W | 2936 hCoV-19/England/QEUH-BC3C64/2020\|EPI_ISL_703936\|20 | ···················· | ···················· | ····················· | ···················· | ···················· | ···················· |
| W | 2937 hCoV-19/England/QEUH-BC37CD/2020\|EPI_ISL_703938\|20 | ···················· | ···················· | ····················· | ···················· | ···················· | ···················· |
| W | 2938 hCoV-19/England/QEUH-BC3C37/2020\|EPI_ISL_703939\|20 | ···················· | ···················· | ····················· | ···················· | ···················· | ···················· |
| W | 2939 hCoV-19/England/QEUH-BC3815/2020\|EPI_ISL_703941\|20 | ···················· | ···················· | ····················· | ···················· | ···················· | ···················· |
| W | 2940 hCoV-19/England/QEUH-BCB482/2020\|EPI_ISL_703959\|20 | ···················· | ···················· | ····················· | ···················· | ···················· | ···················· |
| W | 2941 hCoV-19/England/QEUH-BCC630/2020\|EPI_ISL_703960\|20 | ···················· | ···················· | ····················· | ···················· | ···················· | ···················· |
| W | 2942 hCoV-19/England/QEUH-BCBFDC/2020\|EPI_ISL_703962\|20 | ···················· | ···················· | ····················· | ···················· | ···················· | ···················· |
| W | 2943 hCoV-19/England/QEUH-BC3B2B/2020\|EPI_ISL_703964\|20 | ···················· | ···················· | ····················· | ···················· | ···················· | ···················· |
| W | 2944 hCoV-19/England/QEUH-BCB67D/2020\|EPI_ISL_703965\|20 | ···················· | ···················· | ····················· | ···················· | ···················· | ···················· |
| W | 2945 hCoV-19/England/QEUH-BC3DCB/2020\|EPI_ISL_703967\|20 | ···················· | ···················· | ····················· | ···················· | ···················· | ···················· |
| W | 2946 hCoV-19/England/QEUH-BC3AD3/2020\|EPI_ISL_703972\|20 | ···················· | ···················· | ····················· | ···················· | ···················· | ···················· |
| W | 2947 hCoV-19/England/QEUH-BCB929/2020\|EPI_ISL_703973\|20 | ···················· | ···················· | ····················· | ···················· | ···················· | ···················· |
| W | 2948 hCoV-19/England/QEUH-BC3D8F/2020\|EPI_ISL_703975\|20 | ···················· | ···················· | ····················· | ···················· | ···················· | ···················· |
| W | 2949 hCoV-19/England/QEUH-BCB9A1/2020\|EPI_ISL_703976\|20 | ···················· | ···················· | ····················· | ···················· | ···················· | ···················· |
| W | 2950 hCoV-19/England/QEUH-BC4B39/2020\|EPI_ISL_703978\|20 | ···················· | ···················· | ····················· | ···················· | ···················· | ···················· |
| W | 2951 hCoV-19/England/MILK-B3BE34/2020\|EPI_ISL_706100\|20 | ···················· | ···················· | ····················· | ···················· | ···················· | ···················· |
| W | 2952 hCoV-19/England/MILK-B3BE61/2020\|EPI_ISL_706101\|20 | ···················· | ···················· | ····················· | ···················· | ···················· | ···················· |
| W | 2953 hCoV-19/England/MILK-B3BE70/2020\|EPI_ISL_706102\|20 | ···················· | ···················· | ····················· | ···················· | ···················· | ···················· |
| W | 2954 hCoV-19/England/MILK-B3BE8F/2020\|EPI_ISL_706103\|20 | ···················· | ···················· | ····················· | ···················· | ···················· | ···················· |
| W | 2955 hCoV-19/England/MILK-B3BEAD/2020\|EPI_ISL_706104\|20 | ···················· | ···················· | ····················· | ···················· | ····T·T············· | ···················· |
| W | 2956 hCoV-19/England/MILK-B3BEBC/2020\|EPI_ISL_706105\|20 | ···················· | ···················· | ····················· | ···················· | ···················· | ···················· |
| W | 2957 hCoV-19/England/MILK-B3BECB/2020\|EPI_ISL_706106\|20 | ···················· | ·A·················· | ····················· | ···················· | ···················· | ···A················ |
| W | 2958 hCoV-19/England/MILK-B3BEDA/2020\|EPI_ISL_706107\|20 | ···················· | ···················· | ····················· | ···················· | ···················· | ···················· |
| W | 2959 hCoV-19/England/MILK-B3BF31/2020\|EPI_ISL_706108\|20 | ···················· | ···················· | ····················· | ···················· | ···················· | ···················· |
| W | 2960 hCoV-19/England/MILK-B3BF8C/2020\|EPI_ISL_706109\|20 | ···················· | ···················· | ····················· | ···················· | ···················· | ···················· |
| W | 2961 hCoV-19/England/MILK-B3BF9B/2020\|EPI_ISL_706110\|20 | ···················· | ···················· | ····················· | ···················· | ···················· | ···················· |
| W | 2962 hCoV-19/England/MILK-B3BFB9/2020\|EPI_ISL_706111\|20 | ···················· | ···················· | ····················· | ···················· | ···················· | ···················· |
| W | 2963 hCoV-19/England/MILK-B3BFC8/2020\|EPI_ISL_706112\|20 | ···················· | ···················· | ····················· | ···················· | ···················· | ···················· |
| W | 2964 hCoV-19/England/MILK-B3C000/2020\|EPI_ISL_706113\|20 | ···················· | ···················· | ····················· | ···················· | ···················· | ···················· |
| W | 2965 hCoV-19/England/MILK-B3C02E/2020\|EPI_ISL_706114\|20 | ···················· | ···················· | ····················· | ···················· | ···················· | ···················· |
| W | 2966 hCoV-19/England/MILK-B3C04C/2020\|EPI_ISL_706115\|20 | ···················· | ···················· | ····················· | ···················· | ···················· | ···················· |
| W | 2967 hCoV-19/England/MILK-B3C06A/2020\|EPI_ISL_706116\|20 | ···················· | ···················· | ····················· | ···················· | ···················· | ···················· |
| W | 2968 hCoV-19/England/MILK-B3C0A6/2020\|EPI_ISL_706117\|20 | ···················· | ···················· | ····················· | ···················· | ···················· | ···················· |
| W | 2969 hCoV-19/England/MILK-B3C0B5/2020\|EPI_ISL_706118\|20 | ···················· | ···················· | ····················· | ···················· | ···················· | ···················· |
| W | 2970 hCoV-19/England/MILK-B3C0C4/2020\|EPI_ISL_706119\|20 | ···················· | ···················· | ····················· | ···················· | ···················· | ···················· |
| W | 2971 hCoV-19/England/MILK-B3C0F1/2020\|EPI_ISL_706120\|20 | ···················· | ···················· | ····················· | ···················· | ···················· | ···················· |

W 2972 hCoV-19/England/MILK-B3C11C/2020|EPI_ISL_706121|20 ···················· ···················· ···················· ···················· ···················· ····················
W 2973 hCoV-19/England/MILK-B3C158/2020|EPI_ISL_706122|20 ···················· ···················· ···················· ···················· ···················· ····················
W 2974 hCoV-19/England/MILK-B3C167/2020|EPI_ISL_706123|20 ···················· ···················· ···················· ···················· ···················· ····················
W 2975 hCoV-19/Wales/MILK-B3C176/2020|EPI_ISL_706124|2020 ···················· ···················· ···················· ···················· ···················· ····················
W 2976 hCoV-19/England/MILK-B3C194/2020|EPI_ISL_706125|20 ···················· ···················· ···················· ···················· ···················· ····················
W 2977 hCoV-19/England/MILK-B3C1A3/2020|EPI_ISL_706126|20 ···················· ···················· ···················· ···················· ···················· ····················
W 2978 hCoV-19/England/MILK-B3C1B2/2020|EPI_ISL_706127|20 ···················· ···················· ···················· ···················· ···················· ····················
W 2979 hCoV-19/England/MILK-B3C1C1/2020|EPI_ISL_706128|20 ···················· ···················· ···················· ···················· ···················· ····················
W 2980 hCoV-19/England/MILK-B3C1D0/2020|EPI_ISL_706129|20 ···················· ···················· ···················· ···················· ···················· ····················
W 2981 hCoV-19/England/MILK-B3C1EF/2020|EPI_ISL_706130|20 ···················· ··············C····· ···················· ···················· ···················· ····················
W 2982 hCoV-19/England/MILK-B3C1FE/2020|EPI_ISL_706131|20 ···················· ···················· ···················· ···················· ······T············· ····················
W 2983 hCoV-19/England/MILK-B3C219/2020|EPI_ISL_706132|20 ···················· ···················· ···················· ···················· ···················· ····················
W 2984 hCoV-19/England/MILK-B3C246/2020|EPI_ISL_706133|20 ···················· ···················· ···················· ···················· ···················· ····················
W 2985 hCoV-19/England/MILK-B3C255/2020|EPI_ISL_706134|20 ···················· ···················· ···················· ···················· ···················· ····················
W 2986 hCoV-19/England/MILK-B3C307/2020|EPI_ISL_706135|20 ···················· ···················· ···················· ···················· ···················· ····················
W 2987 hCoV-19/England/MILK-B3C361/2020|EPI_ISL_706136|20 ···················· ···················· ···················· ···················· ···················· ····················
W 2988 hCoV-19/England/MILK-B3C370/2020|EPI_ISL_706137|20 ···················· ···················· ···················· ···················· ···················· ····················
W 2989 hCoV-19/England/MILK-B3C38F/2020|EPI_ISL_706138|20 ···················· ···················· ···················· ···················· ···················· ····················
W 2990 hCoV-19/England/MILK-B3C3AD/2020|EPI_ISL_706139|20 ···················· ···················· ···················· ···················· ···················· ····················
W 2991 hCoV-19/England/MILK-B3C3DA/2020|EPI_ISL_706140|20 ···················· ···················· ···················· ···················· ···················· ····················
W 2992 hCoV-19/England/MILK-B3C3E9/2020|EPI_ISL_706141|20 ···················· ···················· ···················· ···················· ···················· ····················
W 2993 hCoV-19/England/MILK-B3C3F8/2020|EPI_ISL_706142|20 ···················· ···················· ···················· ···················· ···················· ····················
W 2994 hCoV-19/England/MILK-B3C422/2020|EPI_ISL_706143|20 ···················· ···················· ···················· ···················· ···················· ····················
W 2995 hCoV-19/England/MILK-B3C431/2020|EPI_ISL_706144|20 ···················· ···················· ···················· ···················· ···················· ····················
W 2996 hCoV-19/England/MILK-B3C45F/2020|EPI_ISL_706145|20 ···················· ···················· ···················· ···················· ···················· ····················
W 2997 hCoV-19/England/MILK-B3C48C/2020|EPI_ISL_706146|20 ···················· ···················· ···················· ···················· ···················· ····················
W 2998 hCoV-19/England/MILK-B3C49B/2020|EPI_ISL_706147|20 ···················· ···················· ···················· ···················· ···················· ····················
W 2999 hCoV-19/England/MILK-B3C4AA/2020|EPI_ISL_706148|20 ···················· ···················· ···················· ···················· ···················· ····················
W 3000 hCoV-19/England/MILK-B3C4B9/2020|EPI_ISL_706149|20 ···················· ···················· ···················· ···················· ···················· ····················
W 3001 hCoV-19/England/MILK-B3C4C8/2020|EPI_ISL_706150|20 ···················· ···················· ···················· ···················· ···················· ····················
W 3002 hCoV-19/England/MILK-B3C4D7/2020|EPI_ISL_706151|20 ···················· ···················· ···················· ···················· ···················· ····················
W 3003 hCoV-19/England/MILK-B3C695/2020|EPI_ISL_706152|20 ···················· ···················· ···················· ···················· ···················· ····················
W 3004 hCoV-19/England/MILK-B3C6A4/2020|EPI_ISL_706153|20 ···················· ···················· ···················· ···················· ···················· ····················
W 3005 hCoV-19/England/MILK-B3C6E0/2020|EPI_ISL_706154|20 ···················· ·A·················· ···················· ···················· ···················· ···A················
W 3006 hCoV-19/England/MILK-B3C70B/2020|EPI_ISL_706155|20 ···················· ···················· ···················· ···················· ···················· ····················
W 3007 hCoV-19/England/MILK-B3C765/2020|EPI_ISL_706156|20 ···················· ···················· ···················· ···················· ······T············· ····················
W 3008 hCoV-19/England/MILK-B3C792/2020|EPI_ISL_706157|20 ···················· ···················· ···················· ···················· ···················· ····················
W 3009 hCoV-19/England/MILK-B3C7A1/2020|EPI_ISL_706158|20 ···················· ···················· ···················· ···················· ···················· ····················
W 3010 hCoV-19/England/MILK-B3C7FC/2020|EPI_ISL_706159|20 ···················· ···················· ···················· ···················· ···················· ····················
W 3011 hCoV-19/England/MILK-B3C817/2020|EPI_ISL_706160|20 ···················· ···················· ···················· ···················· ···················· ····················
W 3012 hCoV-19/England/MILK-B3C835/2020|EPI_ISL_706161|20 ···················· ·A·················· ···················· ···················· ···················· ···A················
W 3013 hCoV-19/England/MILK-B3C853/2020|EPI_ISL_706162|20 ···················· ···················· ···················· ···················· ···················· ····················
W 3014 hCoV-19/England/MILK-B3C880/2020|EPI_ISL_706163|20 ···················· ···················· ···················· ···················· ···················· ····················
W 3015 hCoV-19/England/MILK-B3C89F/2020|EPI_ISL_706164|20 ···················· ···················· ···················· ···················· ···················· ····················
W 3016 hCoV-19/England/MILK-B3C8AE/2020|EPI_ISL_706165|20 ···················· ···················· ···················· ···················· ···················· ····················
W 3017 hCoV-19/England/MILK-B3C8CC/2020|EPI_ISL_706166|20 ···················· ···················· ···················· ···················· ···················· ····················
W 3018 hCoV-19/England/MILK-B3C8DB/2020|EPI_ISL_706167|20 ···················· ···················· ···················· ···················· ···················· ····················
W 3019 hCoV-19/England/MILK-B3C923/2020|EPI_ISL_706168|20 ···················· ···················· ···················· ···················· ···················· ····················
W 3020 hCoV-19/England/MILK-B3C932/2020|EPI_ISL_706169|20 ···················· ···················· ···················· ···················· ···················· ····················
W 3021 hCoV-19/England/MILK-B3C941/2020|EPI_ISL_706170|20 ···················· ···················· ···················· ···················· ···················· ····················
W 3022 hCoV-19/England/MILK-B3C97E/2020|EPI_ISL_706171|20 ···················· ···················· ···················· ···················· ···················· ····················
W 3023 hCoV-19/England/MILK-B3C99C/2020|EPI_ISL_706172|20 ···················· ···················· ···················· ···················· ···················· ····················
W 3024 hCoV-19/England/MILK-B3C9AB/2020|EPI_ISL_706173|20 ···················· ···················· ···················· ···················· ···················· ····················
W 3025 hCoV-19/England/MILK-B3C9BA/2020|EPI_ISL_706174|20 ···················· ···················· ···················· ···················· ···················· ····················
W 3026 hCoV-19/England/MILK-B3C9C9/2020|EPI_ISL_706175|20 ···················· ···················· ···················· ···················· ···················· ····················
W 3027 hCoV-19/England/MILK-B3C9D8/2020|EPI_ISL_706176|20 ···················· ···················· ···················· ···················· ···················· ····················
W 3028 hCoV-19/England/MILK-B3C9F6/2020|EPI_ISL_706177|20 ···················· ···················· ···················· ···················· ···················· ····················
W 3029 hCoV-19/England/MILK-B3CA02/2020|EPI_ISL_706178|20 ···················· ···················· ···················· ···················· ···················· ····················
W 3030 hCoV-19/England/MILK-B3CA11/2020|EPI_ISL_706179|20 ···················· ···················· ···················· ···················· ···················· ····················
W 3031 hCoV-19/England/MILK-B3CA4E/2020|EPI_ISL_706180|20 ···················· ···················· ···················· ···················· ···················· ····················
W 3032 hCoV-19/England/MILK-B3CA7B/2020|EPI_ISL_706181|20 ···················· ···················· ···················· ···················· ···················· ····················
W 3033 hCoV-19/England/MILK-B3CA99/2020|EPI_ISL_706182|20 ···················· ···················· ···················· ···················· ···················· ····················
W 3034 hCoV-19/England/MILK-B3CAA8/2020|EPI_ISL_706183|20 ···················· ···················· ···················· ···················· ···················· ····················
W 3035 hCoV-19/England/MILK-B3CAC6/2020|EPI_ISL_706184|20 ···················· ···················· ···················· ···················· ···················· ····················
W 3036 hCoV-19/England/MILK-B3CB2D/2020|EPI_ISL_706185|20 ···················· ·A····A············· ···················· ···················· ···················· ····················
W 3037 hCoV-19/England/MILK-B3CB4B/2020|EPI_ISL_706186|20 ···················· ···················· ···················· ···················· ···················· ····················
W 3038 hCoV-19/England/MILK-B3CB5A/2020|EPI_ISL_706187|20 ···················· ·A·················· ···················· ···················· ···················· ····················
W 3039 hCoV-19/England/MILK-B3CB78/2020|EPI_ISL_706188|20 ···················· ···················· ···················· ···················· ···················· ····················
W 3040 hCoV-19/England/MILK-B3CB96/2020|EPI_ISL_706189|20 ···················· ···················· ···················· ···················· ···················· ····················
W 3041 hCoV-19/England/MILK-B3CBD2/2020|EPI_ISL_706190|20 ···················· ···················· ···················· ···················· ···················· ····················
W 3042 hCoV-19/England/MILK-B3CBE1/2020|EPI_ISL_706191|20 ···················· ···················· ···················· ···················· ···················· ····················
W 3043 hCoV-19/England/MILK-B3CD27/2020|EPI_ISL_706192|20 ···················· ···················· ···················· ···················· ···················· ····················
W 3044 hCoV-19/England/MILK-B3CD36/2020|EPI_ISL_706193|20 ···················· ···················· ···················· ···················· ···················· ····················
W 3045 hCoV-19/England/MILK-B3CD45/2020|EPI_ISL_706195|20 ···················· ···················· ···················· ···················· ······T············· ····················
W 3046 hCoV-19/England/MILK-B3CD63/2020|EPI_ISL_706196|20 ···················· ···················· ···················· ···················· ···················· ····················
W 3047 hCoV-19/England/MILK-B3CD72/2020|EPI_ISL_706197|20 ···················· ···················· ···················· ···················· ···················· ····················
W 3048 hCoV-19/England/MILK-B3CD90/2020|EPI_ISL_706198|20 ···················· ···················· ···················· ···················· ···················· ····················
W 3049 hCoV-19/England/MILK-B3CDEB/2020|EPI_ISL_706199|20 ···················· ···················· ···················· ···················· ······T············· ····················
W 3050 hCoV-19/Thailand/Bangkok-PT387/2020|EPI_ISL_708721 ···················· ···················· ···················· ···················· ···················· ····················
W 3051 hCoV-19/Thailand/Bangkok-PT388/2020|EPI_ISL_708722 ···················· ···················· ···················· ···················· ···················· ····················

| | | | | | | | |
|---|---|---|---|---|---|---|---|
| W | 3052 hCoV-19/Thailand/Phuket-SQ-AS23/2020\|EPI_ISL_70872 | ···················· | ···················· | ···················· | ···················· | ···················· | ···················· |
| W | 3053 hCoV-19/USA/CA-LACPHL-AE00032/2020\|EPI_ISL_708724\| | ···················· | ····T··············· | ···················· | ···················· | ···················· | ···················· |
| W | 3054 hCoV-19/USA/CA-LACPHL-AE00033/2020\|EPI_ISL_708725\| | ···················· | ····T··············· | ···················· | ···················· | ···················· | ···················· |
| W | 3055 hCoV-19/USA/CA-LACPHL-AE00034/2020\|EPI_ISL_708726\| | ···················· | ···················· | ···················· | ···················· | ···················· | ···················· |
| W | 3056 hCoV-19/USA/CA-LACPHL-AE00035/2020\|EPI_ISL_708727\| | ···················· | ····T··············· | ···················· | ···················· | ···················· | ···················· |
| W | 3057 hCoV-19/USA/CA-LACPHL-AE00037/2020\|EPI_ISL_708729\| | ···················· | ···················· | ···················· | ···················· | ···················· | ···················· |
| W | 3058 hCoV-19/USA/CA-LACPHL-AE00038/2020\|EPI_ISL_708730\| | ···················· | ····T··············· | ···················· | ···················· | ···················· | ···················· |
| W | 3059 hCoV-19/USA/CA-LACPHL-AE00040/2020\|EPI_ISL_708732\| | ···················· | ····T··············· | ···················· | ···················· | ···················· | ···················· |
| W | 3060 hCoV-19/USA/CA-LACPHL-AE00041/2020\|EPI_ISL_708733\| | ···················· | ····T··············· | ···················· | ···················· | ···················· | ···················· |
| W | 3061 hCoV-19/USA/CA-LACPHL-AE00042/2020\|EPI_ISL_708734\| | ···················· | ···················· | ···················· | ···················· | ···················· | ···················· |
| W | 3062 hCoV-19/Thailand/Phuket-SQ-JS21/2020\|EPI_ISL_70873 | ···················· | ···················· | ···················· | ···················· | ···················· | ···················· |
| W | 3063 hCoV-19/Thailand/Bangkok-025900/2020\|EPI_ISL_70873 | ···················· | ···················· | ···················· | ···················· | ···················· | ···················· |
| W | 3064 hCoV-19/Thailand/Chonburi-SQ-KC0001/2020\|EPI_ISL_7 | ···················· | ···················· | ···················· | ···················· | ···················· | ···················· |
| W | 3065 hCoV-19/Australia/WA444/2020\|EPI_ISL_708738\|2020-0 | ···················· | ···················· | ···················· | ···················· | ···················· | ···················· |
| W | 3066 hCoV-19/Australia/WA445/2020\|EPI_ISL_708739\|2020-0 | ···················· | ···················· | ···················· | ···················· | ···················· | ···················· |
| W | 3067 hCoV-19/Australia/WA495/2020\|EPI_ISL_708789\|2020-1 | ···················· | ···················· | ···················· | ···················· | ···················· | ···················· |
| W | 3068 hCoV-19/Australia/WA496/2020\|EPI_ISL_708790\|2020-1 | ···················· | ···················· | ···················· | ···················· | ···················· | ···················· |
| W | 3069 hCoV-19/Australia/WA499/2020\|EPI_ISL_708793\|2020-1 | ···················· | ···················· | ···················· | ···················· | ···················· | ···················· |
| W | 3070 hCoV-19/Thailand/Chonburi-SQ-NT0002/2020\|EPI_ISL_7 | ···················· | ···················· | ···················· | ···················· | ···················· | ···················· |
| W | 3071 hCoV-19/Australia/NSW1219/2020\|EPI_ISL_708798\|2020 | ···················· | ···················· | ···················· | ···················· | ···················· | ···················· |
| W | 3072 hCoV-19/Australia/NSW1220/2020\|EPI_ISL_708799\|2020 | ···················· | ···················· | ···················· | ···················· | ···················· | ···················· |
| W | 3073 hCoV-19/USA/AZ-TG491661/2020\|EPI_ISL_695205\|2020-0 | ···················· | ·A·················· | ···················· | ···················· | ···················· | ···················· |
| W | 3074 hCoV-19/USA/AZ-TG491664/2020\|EPI_ISL_695206\|2020-0 | ···················· | ·A·················· | ···················· | ···················· | ···················· | ···················· |
| W | 3075 hCoV-19/USA/AZ-TG491665/2020\|EPI_ISL_695207\|2020-0 | ···················· | ·A·················· | ···················· | ···················· | ···················· | ···················· |
| W | 3076 hCoV-19/USA/AZ-TG491673/2020\|EPI_ISL_695209\|2020-0 | ···················· | ·············A······ | ···················· | ···················· | ···················· | ···················· |
| W | 3077 hCoV-19/USA/AZ-TG491705/2020\|EPI_ISL_695220\|2020-0 | ···················· | ·A·················· | ···················· | ···················· | ···················· | ···················· |
| W | 3078 hCoV-19/USA/AZ-TG491707/2020\|EPI_ISL_695221\|2020-0 | ···················· | ·A·················· | ···················· | ···················· | ···················· | ···················· |
| W | 3079 hCoV-19/USA/AZ-TG491713/2020\|EPI_ISL_695224\|2020-0 | ···················· | ·A·················· | ···················· | ···················· | ···················· | ···················· |
| W | 3080 hCoV-19/USA/AZ-TG493823/2020\|EPI_ISL_695237\|2020-0 | ···················· | ···················· | ···················· | ···················· | ···················· | ···················· |
| W | 3081 hCoV-19/USA/AZ-TG494588/2020\|EPI_ISL_695247\|2020-0 | ···················· | ···················· | ···················· | ···················· | ···················· | ···················· |
| W | 3082 hCoV-19/USA/AZ-TG494613/2020\|EPI_ISL_695249\|2020-0 | ···················· | ···················· | ···················· | ···················· | ···················· | ···················· |
| W | 3083 hCoV-19/USA/AZ-TG494830/2020\|EPI_ISL_695251\|2020-0 | ···················· | ·A·················· | ···················· | ···················· | ···················· | ···················· |
| W | 3084 hCoV-19/USA/AZ-TG494842/2020\|EPI_ISL_695252\|2020-0 | ···················· | ·A·················· | ···················· | ···················· | ···················· | ···················· |
| W | 3085 hCoV-19/USA/AZ-TG494849/2020\|EPI_ISL_695255\|2020-0 | ···················· | ···················· | ···················· | ···················· | ···················· | ···················· |
| W | 3086 hCoV-19/USA/AZ-TG494864/2020\|EPI_ISL_695256\|2020-0 | ···················· | ···················· | ···················· | ···················· | ···················· | ···················· |
| W | 3087 hCoV-19/USA/AZ-TG495227/2020\|EPI_ISL_695257\|2020-0 | ···················· | ·A·················· | ···················· | ···················· | ···················· | ···················· |
| W | 3088 hCoV-19/USA/AZ-TG495452/2020\|EPI_ISL_695260\|2020-0 | ···················· | ···················· | ···················· | ···················· | ···················· | ···················· |
| W | 3089 hCoV-19/USA/AZ-TG495769/2020\|EPI_ISL_695262\|2020-0 | ···················· | ···················· | ···················· | ···················· | ···················· | ···················· |
| W | 3090 hCoV-19/USA/AZ-TG495770/2020\|EPI_ISL_695263\|2020-0 | ···················· | ···················· | ···················· | ···················· | ···················· | ···················· |
| W | 3091 hCoV-19/USA/AZ-TG497217/2020\|EPI_ISL_695265\|2020-0 | ···················· | ·A·················· | ···················· | ···················· | ···················· | ···················· |
| W | 3092 hCoV-19/USA/AZ-TG498743/2020\|EPI_ISL_695269\|2020-0 | ···················· | ·A·················· | ···················· | ···················· | ···················· | ···················· |
| W | 3093 hCoV-19/USA/AZ-TG501912/2020\|EPI_ISL_695276\|2020-0 | ···················· | ·A·················· | ···················· | ···················· | ···················· | ···················· |
| W | 3094 hCoV-19/USA/AZ-TG501918/2020\|EPI_ISL_695277\|2020-0 | ···················· | ·A·················· | ···················· | ···················· | ···················· | ···················· |
| W | 3095 hCoV-19/USA/AZ-TG501925/2020\|EPI_ISL_695278\|2020-0 | ···················· | ·A·················· | ···················· | ···················· | ···················· | ···················· |
| W | 3096 hCoV-19/USA/AZ-TG503851/2020\|EPI_ISL_695290\|2020-0 | ···················· | ···················· | ···················· | ···················· | ···················· | ···················· |
| W | 3097 hCoV-19/USA/AZ-TG505110/2020\|EPI_ISL_695293\|2020-0 | ···················· | ···················· | ···················· | ···················· | ···················· | ···················· |
| W | 3098 hCoV-19/USA/AZ-TG505113/2020\|EPI_ISL_695294\|2020-0 | ···················· | ·A·················· | ···················· | ···················· | ···················· | ···················· |
| W | 3099 hCoV-19/USA/AZ-TG507253/2020\|EPI_ISL_695296\|2020-0 | ···················· | ·A·················· | ···················· | ···················· | ···················· | ···················· |
| W | 3100 hCoV-19/USA/AZ-TG507341/2020\|EPI_ISL_695297\|2020-0 | ···················· | ···················· | ···················· | ···················· | ···················· | ···················· |
| W | 3101 hCoV-19/India/MH-ACTREC-351/2020\|EPI_ISL_700000\|20 | ···················· | ···················· | ···················· | ···················· | ···················· | ···················· |
| W | 3102 hCoV-19/India/MP-ACTREC-352/2020\|EPI_ISL_700001\|20 | ···················· | ···················· | ···················· | ···················· | ···················· | ···················· |
| W | 3103 hCoV-19/India/MH-ACTREC-353/2020\|EPI_ISL_700002\|20 | ···················· | ···················· | ···················· | ···················· | ···················· | ···················· |
| W | 3104 hCoV-19/India/MH-ACTREC-354/2020\|EPI_ISL_700003\|20 | ···················· | ···················· | ···················· | ···················· | ···················· | ···················· |
| W | 3105 hCoV-19/India/MH-ACTREC-355/2020\|EPI_ISL_700004\|20 | ···················· | ···················· | ···················· | ···················· | ···················· | ···················· |
| W | 3106 hCoV-19/India/MH-ACTREC-356/2020\|EPI_ISL_700005\|20 | ···················· | ···················· | ···················· | ···················· | ···················· | ···················· |
| W | 3107 hCoV-19/India/MH-ACTREC-357/2020\|EPI_ISL_700006\|20 | ···················· | ···················· | ···················· | ···················· | ···················· | ···················· |
| W | 3108 hCoV-19/India/MH-ACTREC-358/2020\|EPI_ISL_700007\|20 | ···················· | ···················· | ···················· | ···················· | ···················· | ···················· |
| W | 3109 hCoV-19/India/MH-ACTREC-359/2020\|EPI_ISL_700008\|20 | ···················· | ···················· | ···················· | ···················· | ···················· | ···················· |
| W | 3110 hCoV-19/India/MH-ACTREC-360/2020\|EPI_ISL_700009\|20 | ···················· | ···················· | ···················· | ···················· | ···················· | ···················· |
| W | 3111 hCoV-19/India/MH-ACTREC-361/2020\|EPI_ISL_700010\|20 | ···················· | ···················· | ···················· | ···················· | ···················· | ···················· |
| W | 3112 hCoV-19/India/MH-ACTREC-363/2020\|EPI_ISL_700012\|20 | ···················· | ···················· | ···················· | ···················· | ···················· | ···················· |
| W | 3113 hCoV-19/India/MH-ACTREC-364/2020\|EPI_ISL_700013\|20 | ···················· | ···················· | ···················· | ···················· | ···················· | ···················· |
| W | 3114 hCoV-19/India/MH-ACTREC-365/2020\|EPI_ISL_700014\|20 | ···················· | ···················· | ···················· | ···················· | ···················· | ···················· |
| W | 3115 hCoV-19/India/MH-ACTREC-367/2020\|EPI_ISL_700016\|20 | ···················· | ···················· | ···················· | ···················· | ···················· | ···················· |
| W | 3116 hCoV-19/India/MH-ACTREC-368/2020\|EPI_ISL_700017\|20 | ···················· | ···················· | ···················· | ···················· | ···················· | ···················· |
| W | 3117 hCoV-19/India/MH-ACTREC-370/2020\|EPI_ISL_700019\|20 | ···················· | ···················· | ···················· | ···················· | ···················· | ···················· |
| W | 3119 hCoV-19/India/MH-ACTREC-372/2020\|EPI_ISL_700021\|20 | ···················· | ···················· | ···················· | ···················· | ···················· | ···················· |
| W | 3120 hCoV-19/India/MH-ACTREC-373/2020\|EPI_ISL_700022\|20 | ···················· | ···················· | ···················· | ···················· | ···················· | ···················· |
| W | 3121 hCoV-19/India/MH-ACTREC-374/2020\|EPI_ISL_700023\|20 | ···················· | ···················· | ···················· | ···················· | ···················· | ···················· |
| W | 3122 hCoV-19/India/MH-ACTREC-375/2020\|EPI_ISL_700024\|20 | ···················· | ····A··············· | ···················· | ···················· | ···················· | ···················· |
| W | 3123 hCoV-19/India/MH-ACTREC-376/2020\|EPI_ISL_700025\|20 | ···················· | ····A··············· | ···················· | ···················· | ···················· | ···················· |
| W | 3124 hCoV-19/India/MH-ACTREC-377/2020\|EPI_ISL_700026\|20 | ···················· | ···················· | ···················· | ···················· | ···················· | ···················· |
| W | 3125 hCoV-19/India/MH-ACTREC-378/2020\|EPI_ISL_700027\|20 | ···················· | ···················· | ···················· | ···················· | ···················· | ···················· |
| W | 3126 hCoV-19/India/MH-ACTREC-379/2020\|EPI_ISL_700028\|20 | ···················· | ···················· | ···················· | ···················· | ···················· | ···················· |
| W | 3127 hCoV-19/India/MH-ACTREC-380/2020\|EPI_ISL_700029\|20 | ···················· | ···················· | ···················· | ···················· | ···················· | ···················· |
| W | 3128 hCoV-19/India/MH-ACTREC-381/2020\|EPI_ISL_700030\|20 | ···················· | ···················· | ···················· | ···················· | ···················· | ···················· |
| W | 3129 hCoV-19/India/MH-ACTREC-382/2020\|EPI_ISL_700031\|20 | ···················· | ···················· | ···················· | ···················· | ···················· | ···················· |
| W | 3130 hCoV-19/India/MH-ACTREC-383/2020\|EPI_ISL_700032\|20 | ···················· | ···················· | ···················· | ···················· | ···················· | ···················· |
| W | 3132 hCoV-19/India/MH-ACTREC-385/2020\|EPI_ISL_700034\|20 | ···················· | ···················· | ···················· | ···················· | ···················· | ···················· |
| W | 3133 hCoV-19/India/MH-ACTREC-386/2020\|EPI_ISL_700035\|20 | ···················· | ···················· | ···················· | ···················· | ···················· | ···················· |

| | | | | | | | | |
|---|---|---|---|---|---|---|---|---|
| W | 3134 | hCoV-19/India/MH-ACTREC-387/2020\|EPI_ISL_700036\|20 | ···················· | ···················· | ····················· | ···················· | ···················· | ···················· |
| W | 3135 | hCoV-19/India/MH-ACTREC-388/2020\|EPI_ISL_700037\|20 | ···················· | ···················· | ····················· | ···················· | ···················· | ···················· |
| W | 3136 | hCoV-19/India/MH-ACTREC-389/2020\|EPI_ISL_700038\|20 | ···················· | ···················· | ····················· | ···················· | ···················· | ···················· |
| W | 3137 | hCoV-19/India/MH-ACTREC-390/2020\|EPI_ISL_700039\|20 | ···················· | ···················· | ····················· | ···················· | ···················· | ···················· |
| W | 3138 | hCoV-19/India/MH-ACTREC-393/2020\|EPI_ISL_700042\|20 | ···················· | ···················· | ····················· | ···················· | ···················· | ···················· |
| W | 3139 | hCoV-19/India/MH-ACTREC-394/2020\|EPI_ISL_700043\|20 | ···················· | ···················· | ····················· | ···················· | ···················· | ···················· |
| W | 3140 | hCoV-19/India/MH-ACTREC-395/2020\|EPI_ISL_700044\|20 | ···················· | ···················· | ····················· | ···················· | ···················· | ···················· |
| W | 3141 | hCoV-19/India/MH-ACTREC-396/2020\|EPI_ISL_700045\|20 | ···················· | ···················· | ····················· | ···················· | ···················· | ···················· |
| W | 3142 | hCoV-19/India/MH-ACTREC-397/2020\|EPI_ISL_700046\|20 | ···················· | ···················· | ····················· | ···················· | ···················· | ···················· |
| W | 3143 | hCoV-19/India/MH-ACTREC-398/2020\|EPI_ISL_700047\|20 | ···················· | ···················· | ····················· | ···················· | ···················· | ···················· |
| W | 3144 | hCoV-19/India/MH-ACTREC-399/2020\|EPI_ISL_700048\|20 | ···················· | ···················· | ····················· | ···················· | ···················· | ···················· |
| W | 3145 | hCoV-19/India/MH-ACTREC-400/2020\|EPI_ISL_700049\|20 | ···················· | ···················· | ····················· | ···················· | ···················· | ···················· |
| W | 3146 | hCoV-19/India/MH-ACTREC-401/2020\|EPI_ISL_700050\|20 | ···················· | ···················· | ····················· | ···················· | ···················· | ···················· |
| W | 3147 | hCoV-19/India/MH-ACTREC-402/2020\|EPI_ISL_700051\|20 | ···················· | ···················· | ····················· | ···················· | ···················· | ···················· |
| W | 3148 | hCoV-19/India/MH-ACTREC-403/2020\|EPI_ISL_700052\|20 | ···················· | ···················· | ····················· | ···················· | ···················· | ···················· |
| W | 3149 | hCoV-19/India/MH-ACTREC-404/2020\|EPI_ISL_700053\|20 | ···················· | ···················· | ····················· | ···················· | ···················· | ···················· |
| W | 3150 | hCoV-19/India/MH-ACTREC-405/2020\|EPI_ISL_700054\|20 | ···················· | ···················· | ····················· | ···················· | ···················· | ···················· |
| W | 3151 | hCoV-19/India/MH-ACTREC-406/2020\|EPI_ISL_700055\|20 | ···················· | ···················· | ····················· | ···················· | ···················· | ···················· |
| W | 3153 | hCoV-19/India/MH-ACTREC-408/2020\|EPI_ISL_700057\|20 | ···················· | ···················· | ····················· | ···················· | ···················· | ···················· |
| W | 3154 | hCoV-19/India/MH-ACTREC-409/2020\|EPI_ISL_700058\|20 | ···················· | ···················· | ····················· | ···················· | ···················· | ···················· |
| W | 3155 | hCoV-19/India/MH-ACTREC-410/2020\|EPI_ISL_700059\|20 | ···················· | ···················· | ····················· | ···················· | ···················· | ···················· |
| W | 3156 | hCoV-19/India/MH-ACTREC-411/2020\|EPI_ISL_700060\|20 | ···················· | ···················· | ····················· | ···················· | ···················· | ···················· |
| W | 3157 | hCoV-19/India/MH-ACTREC-412/2020\|EPI_ISL_700061\|20 | ···················· | ···················· | ····················· | ···················· | ···················· | ···················· |
| W | 3158 | hCoV-19/India/MH-ACTREC-413/2020\|EPI_ISL_700062\|20 | ···················· | ···················· | ····················· | ···················· | ···················· | ···················· |
| W | 3159 | hCoV-19/India/MH-ACTREC-414/2020\|EPI_ISL_700063\|20 | ···················· | ···················· | ····················· | ···················· | ···················· | ···················· |
| W | 3160 | hCoV-19/India/MH-ACTREC-415/2020\|EPI_ISL_700064\|20 | ···················· | ···················· | ····················· | ···················· | ···················· | ···················· |
| W | 3161 | hCoV-19/India/MH-ACTREC-416/2020\|EPI_ISL_700065\|20 | ···················· | ····G··············· | ····················· | ···················· | ···················· | ···················· |
| W | 3162 | hCoV-19/India/MH-ACTREC-417/2020\|EPI_ISL_700066\|20 | ···················· | ···················· | ····················· | ···················· | ···················· | ···················· |
| W | 3163 | hCoV-19/India/MH-ACTREC-418/2020\|EPI_ISL_700067\|20 | ···················· | ···················· | ····················· | ···················· | ···················· | ···················· |
| W | 3164 | hCoV-19/India/MH-ACTREC-419/2020\|EPI_ISL_700068\|20 | ···················· | ······A············· | ····················· | ···················· | ···················· | ···················· |
| W | 3165 | hCoV-19/India/MH-ACTREC-420/2020\|EPI_ISL_700069\|20 | ···················· | ···················· | ····················· | ···················· | ···················· | ···················· |
| W | 3166 | hCoV-19/India/UP-ACTREC-421/2020\|EPI_ISL_700070\|20 | ···················· | ···················· | ····················· | ···················· | ···················· | ···················· |
| W | 3167 | hCoV-19/India/MH-ACTREC-422/2020\|EPI_ISL_700071\|20 | ···················· | ···················· | ····················· | ···················· | ···················· | ···················· |
| W | 3168 | hCoV-19/India/MH-ACTREC-423/2020\|EPI_ISL_700072\|20 | ···················· | ···················· | ····················· | ···················· | ···················· | ···················· |
| W | 3170 | hCoV-19/India/MH-ACTREC-425/2020\|EPI_ISL_700074\|20 | ···················· | ···················· | ····················· | ···················· | ···················· | ···················· |
| W | 3171 | hCoV-19/India/MH-ACTREC-426/2020\|EPI_ISL_700075\|20 | ···················· | ···················· | ····················· | ···················· | ···················· | ···················· |
| W | 3172 | hCoV-19/India/MH-ACTREC-427/2020\|EPI_ISL_700076\|20 | ···················· | ···················· | ····················· | ···················· | ···················· | ···················· |
| W | 3173 | hCoV-19/India/MH-ACTREC-428/2020\|EPI_ISL_700077\|20 | ···················· | ···················· | ····················· | ···················· | ···················· | ···················· |
| W | 3174 | hCoV-19/India/MH-ACTREC-429/2020\|EPI_ISL_700078\|20 | ···················· | ···················· | ····················· | ···················· | ···················· | ···················· |
| W | 3175 | hCoV-19/India/MH-ACTREC-431/2020\|EPI_ISL_700080\|20 | ···················· | ···················· | ····················· | ···················· | ···················· | ···················· |
| W | 3176 | hCoV-19/India/MH-ACTREC-432/2020\|EPI_ISL_700081\|20 | ···················· | ···················· | ····················· | ···················· | ···················· | ···················· |
| W | 3178 | hCoV-19/India/MH-ACTREC-434/2020\|EPI_ISL_700083\|20 | ···················· | ···················· | ····················· | ···················· | ···················· | ···················· |
| W | 3179 | hCoV-19/India/MH-ACTREC-435/2020\|EPI_ISL_700084\|20 | ···················· | ···················· | ····················· | ···················· | ···················· | ···················· |
| W | 3182 | hCoV-19/India/MH-ACTREC-438/2020\|EPI_ISL_700087\|20 | ···················· | ···················· | ····················· | ···················· | ···················· | ···················· |
| W | 3183 | hCoV-19/India/MH-ACTREC-439/2020\|EPI_ISL_700088\|20 | ···················· | ···················· | ····················· | ···················· | ···················· | ···················· |
| W | 3187 | hCoV-19/India/MH-ACTREC-444/2020\|EPI_ISL_700092\|20 | ···················· | ···················· | ····················· | ···················· | ···················· | ···················· |
| W | 3188 | hCoV-19/India/MH-ACTREC-445/2020\|EPI_ISL_700093\|20 | ···················· | ···················· | ····················· | ···················· | ···················· | ···················· |
| W | 3189 | hCoV-19/India/MH-ACTREC-446/2020\|EPI_ISL_700094\|20 | ···················· | ···················· | ····················· | ···················· | ···················· | ···················· |
| W | 3190 | hCoV-19/India/MH-ACTREC-447/2020\|EPI_ISL_700095\|20 | ···················· | ···················· | ····················· | ···················· | ···················· | ···················· |
| W | 3191 | hCoV-19/India/MH-ACTREC-448/2020\|EPI_ISL_700096\|20 | ···················· | ···················· | ····················· | ···················· | ···················· | ···················· |
| W | 3192 | hCoV-19/India/MH-ACTREC-449/2020\|EPI_ISL_700097\|20 | ···················· | ···················· | ····················· | ···················· | ···················· | ···················· |
| W | 3193 | hCoV-19/India/MH-ACTREC-450/2020\|EPI_ISL_700098\|20 | ···················· | ···················· | ····················· | ···················· | ···················· | ···················· |
| W | 3194 | hCoV-19/India/MH-ACTREC-451/2020\|EPI_ISL_700099\|20 | ···················· | ···················· | ····················· | ···················· | ···················· | ···················· |
| W | 3195 | hCoV-19/England/ALDP-BBA91E/2020\|EPI_ISL_702613\|20 | ···················· | ···················· | ····················· | ···················· | ···················· | ···················· |
| W | 3196 | hCoV-19/England/ALDP-BB98D6/2020\|EPI_ISL_702614\|20 | ···················· | ···················· | ····················· | ···················· | ···················· | ···················· |
| W | 3197 | hCoV-19/England/ALDP-BB8C9F/2020\|EPI_ISL_702616\|20 | ···················· | ···················· | ····················· | ···················· | ···················· | ···················· |
| W | 3198 | hCoV-19/England/ALDP-BB5782/2020\|EPI_ISL_702617\|20 | ···················· | ···················· | ····················· | ···················· | ···················· | ···················· |
| W | 3199 | hCoV-19/England/ALDP-BB3450/2020\|EPI_ISL_702619\|20 | ···················· | ···················· | ····················· | ···················· | ···················· | ···················· |
| W | 3200 | hCoV-19/England/ALDP-BB993D/2020\|EPI_ISL_702621\|20 | ···················· | ···················· | ····················· | ···················· | ···················· | ···················· |
| W | 3201 | hCoV-19/England/ALDP-BB98C7/2020\|EPI_ISL_702624\|20 | ···················· | ···················· | ····················· | ···················· | ···················· | ···················· |
| W | 3202 | hCoV-19/England/ALDP-BBACBB/2020\|EPI_ISL_702650\|20 | ···················· | ···················· | ····················· | ···················· | ···················· | ···················· |
| W | 3203 | hCoV-19/England/ALDP-BB5922/2020\|EPI_ISL_702651\|20 | ···················· | ···················· | ····················· | ···················· | ···················· | ···················· |
| W | 3204 | hCoV-19/England/ALDP-BB99B5/2020\|EPI_ISL_702653\|20 | ···················· | ···················· | ····················· | ···················· | ···················· | ···················· |
| W | 3205 | hCoV-19/England/ALDP-BB5755/2020\|EPI_ISL_702657\|20 | ···················· | ···················· | ····················· | ···················· | ···················· | ···················· |
| W | 3206 | hCoV-19/England/ALDP-BB3159/2020\|EPI_ISL_702658\|20 | ···················· | ···················· | ····················· | ···················· | ···················· | ···················· |
| W | 3207 | hCoV-19/England/ALDP-BB8E4E/2020\|EPI_ISL_702660\|20 | ···················· | ···················· | ····················· | ···················· | ···················· | ···················· |
| W | 3208 | hCoV-19/England/ALDP-BB9812/2020\|EPI_ISL_702662\|20 | ···················· | ···················· | ····················· | ···················· | ···················· | ···················· |
| W | 3209 | hCoV-19/England/ALDP-BB312C/2020\|EPI_ISL_702663\|20 | ···················· | ···················· | ····················· | ···················· | ···················· | ···················· |
| W | 3210 | hCoV-19/England/ALDP-BB3274/2020\|EPI_ISL_702665\|20 | ···················· | ···················· | ····················· | ···················· | ···················· | ···················· |
| W | 3211 | hCoV-19/England/ALDP-BB597D/2020\|EPI_ISL_702671\|20 | ···················· | ···················· | ····················· | ···················· | ···················· | ···················· |
| W | 3212 | hCoV-19/England/ALDP-BB8E11/2020\|EPI_ISL_702675\|20 | ···················· | ···················· | ····················· | ···················· | ···················· | ···················· |
| W | 3213 | hCoV-19/England/ALDP-BB99F1/2020\|EPI_ISL_702676\|20 | ···················· | ···················· | ····················· | ···················· | ······T············· | ···················· |
| W | 3214 | hCoV-19/England/ALDP-BB33AE/2020\|EPI_ISL_702678\|20 | ···················· | ···················· | ····················· | ···················· | ···················· | ···················· |
| W | 3215 | hCoV-19/England/ALDP-BB27C1/2020\|EPI_ISL_702680\|20 | ···················· | ···················· | ····················· | ···················· | ···················· | ···················· |
| W | 3216 | hCoV-19/England/ALDP-BB3423/2020\|EPI_ISL_702681\|20 | ···················· | ···················· | ····················· | ···················· | ···················· | ···················· |
| W | 3217 | hCoV-19/England/ALDP-BB2BF2/2020\|EPI_ISL_702683\|20 | ···················· | ···················· | ····················· | ···················· | ···················· | ···················· |
| W | 3218 | hCoV-19/England/ALDP-BB5931/2020\|EPI_ISL_702686\|20 | ···················· | ···················· | ····················· | ···················· | ···················· | ···················· |
| W | 3219 | hCoV-19/England/ALDP-BB52EB/2020\|EPI_ISL_702688\|20 | ···················· | ···················· | ····················· | ···················· | ···A················ | ···················· |
| W | 3220 | hCoV-19/England/ALDP-BB9A85/2020\|EPI_ISL_702689\|20 | ···················· | ···················· | ····················· | ···················· | ···················· | ···················· |
| W | 3221 | hCoV-19/England/ALDP-BB2B7A/2020\|EPI_ISL_702691\|20 | ···················· | ···················· | ····················· | ···················· | ···················· | ···················· |

| | | | | | | | |
|---|---|---|---|---|---|---|---|
| W | 3222 hCoV-19/England/ALDP-BB8F4B/2020\|EPI_ISL_702693\|20 | ···················· | ···················· | ···················· | ···················· | ···················· | ···················· |
| W | 3223 hCoV-19/England/ALDP-BB8E8A/2020\|EPI_ISL_702696\|20 | ···················· | ···················· | ···················· | ···················· | ···················· | ···················· |
| W | 3224 hCoV-19/England/QEUH-BCB992/2020\|EPI_ISL_703943\|20 | ···················· | ···················· | ···················· | ···················· | ···················· | ···················· |
| W | 3225 hCoV-19/England/QEUH-BCBD78/2020\|EPI_ISL_703944\|20 | ···················· | ···················· | ···················· | ···················· | ···················· | ···················· |
| W | 3226 hCoV-19/England/QEUH-BCB9DE/2020\|EPI_ISL_703949\|20 | ···················· | ···················· | ···················· | ···················· | ···················· | ···················· |
| W | 3227 hCoV-19/England/QEUH-BC4AD2/2020\|EPI_ISL_703951\|20 | ···················· | ·A·················· | ···················· | ···················· | ···················· | ···················· |
| W | 3228 hCoV-19/England/QEUH-BCB877/2020\|EPI_ISL_703956\|20 | ···················· | ···················· | ···················· | ···················· | ···················· | ···················· |
| W | 3229 hCoV-19/England/QEUH-BC3D25/2020\|EPI_ISL_703957\|20 | ···················· | ···················· | ···················· | ···················· | ···················· | ···················· |
| W | 3230 hCoV-19/England/QEUH-BCB8E0/2020\|EPI_ISL_703980\|20 | ···················· | ···················· | ···················· | ···················· | ···················· | ···················· |
| W | 3231 hCoV-19/England/QEUH-BCBA26/2020\|EPI_ISL_703981\|20 | ···················· | ···················· | ···················· | ···················· | ···················· | ···················· |
| W | 3232 hCoV-19/England/QEUH-BCC0C9/2020\|EPI_ISL_703983\|20 | ···················· | ···················· | ···················· | ···················· | ···················· | ···················· |
| W | 3233 hCoV-19/England/QEUH-BCC06F/2020\|EPI_ISL_703984\|20 | ···················· | ···················· | ···················· | ···················· | ···················· | ···················· |
| W | 3234 hCoV-19/England/QEUH-BC3736/2020\|EPI_ISL_703986\|20 | ···················· | ···················· | ···················· | ···················· | ···················· | ···················· |
| W | 3235 hCoV-19/England/QEUH-BC3648/2020\|EPI_ISL_703988\|20 | ···················· | ···················· | ···················· | ···················· | ···················· | ···················· |
| W | 3236 hCoV-19/England/QEUH-BCB81D/2020\|EPI_ISL_703991\|20 | ···················· | ···················· | ···················· | ···················· | ···················· | ···················· |
| W | 3237 hCoV-19/England/QEUH-BCC278/2020\|EPI_ISL_703993\|20 | ···················· | ···················· | ···················· | ···················· | ···················· | ···················· |
| W | 3238 hCoV-19/England/QEUH-BC35D2/2020\|EPI_ISL_703994\|20 | ···················· | ···················· | ···················· | ···················· | ···················· | ···················· |
| W | 3239 hCoV-19/England/QEUH-BCC5CA/2020\|EPI_ISL_703996\|20 | ···················· | ···················· | ···················· | ···················· | ···················· | ···················· |
| W | 3240 hCoV-19/England/QEUH-BC3D61/2020\|EPI_ISL_703997\|20 | ···················· | ···················· | ···················· | ···················· | ···················· | ···A················ |
| W | 3241 hCoV-19/England/QEUH-BC3B76/2020\|EPI_ISL_703999\|20 | ···················· | ···················· | ···················· | ···················· | ···················· | ···················· |
| W | 3242 hCoV-19/England/MILK-B3CE51/2020\|EPI_ISL_706200\|20 | ···················· | ···················· | ···················· | ···················· | ···················· | ···················· |
| W | 3243 hCoV-19/England/MILK-B3CEAC/2020\|EPI_ISL_706201\|20 | ···················· | ···················· | ···················· | ···················· | ···················· | ···················· |
| W | 3244 hCoV-19/England/MILK-B3CEBB/2020\|EPI_ISL_706202\|20 | ···················· | ···················· | ···················· | ···················· | ···················· | ···················· |
| W | 3245 hCoV-19/England/MILK-B3CED9/2020\|EPI_ISL_706203\|20 | ···················· | ···················· | ···················· | ···················· | ···················· | ···················· |
| W | 3246 hCoV-19/England/MILK-B3CEE8/2020\|EPI_ISL_706204\|20 | ···················· | ···················· | ···················· | ···················· | ···················· | ···················· |
| W | 3247 hCoV-19/England/MILK-B3CEF7/2020\|EPI_ISL_706205\|20 | ···················· | ···················· | ···················· | ···················· | ···················· | ···················· |
| W | 3248 hCoV-19/England/MILK-B3CF03/2020\|EPI_ISL_706206\|20 | ···················· | ···················· | ···················· | ···················· | ···················· | ···················· |
| W | 3249 hCoV-19/England/MILK-B3CF21/2020\|EPI_ISL_706207\|20 | ···················· | ···················· | ···················· | ···················· | ···················· | ···················· |
| W | 3250 hCoV-19/England/MILK-B3CF4F/2020\|EPI_ISL_706208\|20 | ···················· | ···················· | ···················· | ···················· | ···················· | ···················· |
| W | 3251 hCoV-19/England/MILK-B3CF6D/2020\|EPI_ISL_706209\|20 | ···················· | ···················· | ···················· | ···················· | ···················· | ···················· |
| W | 3252 hCoV-19/England/MILK-B3CF9A/2020\|EPI_ISL_706210\|20 | ···················· | ···················· | ···················· | ···················· | ···················· | ···················· |
| W | 3253 hCoV-19/England/MILK-B3CFB8/2020\|EPI_ISL_706211\|20 | ···················· | ·A·················· | ···················· | ···················· | ···················· | ···················· |
| W | 3254 hCoV-19/England/MILK-B3D02D/2020\|EPI_ISL_706214\|20 | ···················· | ·A·················· | ···················· | ···················· | ···················· | ···A················ |
| W | 3255 hCoV-19/England/MILK-B3D5F1/2020\|EPI_ISL_706230\|20 | ···················· | ···················· | ···················· | ···················· | ···················· | ···················· |
| W | 3256 hCoV-19/England/MILK-B3D649/2020\|EPI_ISL_706231\|20 | ···················· | ···················· | ···················· | ···················· | ···················· | ···················· |
| W | 3257 hCoV-19/England/MILK-B3D658/2020\|EPI_ISL_706232\|20 | ···················· | ···················· | ···················· | ···················· | ···················· | ···················· |
| W | 3258 hCoV-19/England/MILK-B3D755/2020\|EPI_ISL_706233\|20 | ···················· | ···················· | ···················· | ···················· | ···················· | ···················· |
| W | 3259 hCoV-19/England/MILK-B3D782/2020\|EPI_ISL_706234\|20 | ···················· | ···················· | ···················· | ···················· | ···················· | ···A················ |
| W | 3260 hCoV-19/England/MILK-B3D7BF/2020\|EPI_ISL_706235\|20 | ···················· | ···················· | ···················· | ···················· | ···················· | ···················· |
| W | 3261 hCoV-19/England/MILK-B3D7CE/2020\|EPI_ISL_706236\|20 | ···················· | ···················· | ···················· | ···················· | ···················· | ···················· |
| W | 3262 hCoV-19/England/MILK-B3D7DD/2020\|EPI_ISL_706237\|20 | ···················· | ···················· | ···················· | ···················· | ···················· | ···················· |
| W | 3263 hCoV-19/England/MILK-B3D816/2020\|EPI_ISL_706238\|20 | ···················· | ···················· | ···················· | ···················· | ···················· | ···················· |
| W | 3264 hCoV-19/England/MILK-B3D88F/2020\|EPI_ISL_706239\|20 | ···················· | ···················· | ···················· | ···················· | ···················· | ···A················ |
| W | 3265 hCoV-19/England/MILK-B3D8E9/2020\|EPI_ISL_706240\|20 | ···················· | ···················· | ···················· | ···················· | ···················· | ···················· |
| W | 3266 hCoV-19/England/MILK-B3D913/2020\|EPI_ISL_706241\|20 | ···················· | ···················· | ···················· | ···················· | ···················· | ···················· |
| W | 3267 hCoV-19/England/MILK-B3D95F/2020\|EPI_ISL_706242\|20 | ···················· | ···················· | ···················· | ···················· | ···················· | ···················· |
| W | 3268 hCoV-19/England/MILK-B3D96E/2020\|EPI_ISL_706243\|20 | ···················· | ···················· | ···················· | ···················· | ······T············· | ···················· |
| W | 3269 hCoV-19/England/MILK-B3D98C/2020\|EPI_ISL_706244\|20 | ···················· | ···················· | ···················· | ···················· | ···················· | ···················· |
| W | 3270 hCoV-19/England/MILK-B3DBA4/2020\|EPI_ISL_706245\|20 | ···················· | ···················· | ···················· | ···················· | ···················· | ···················· |
| W | 3271 hCoV-19/England/MILK-B3DBC2/2020\|EPI_ISL_706246\|20 | ···················· | ···················· | ···················· | ···················· | ······T············· | ···················· |
| W | 3272 hCoV-19/England/MILK-B3DBD1/2020\|EPI_ISL_706247\|20 | ···················· | ···················· | ···················· | ···················· | ···················· | ···················· |
| W | 3273 hCoV-19/England/MILK-B3DBFF/2020\|EPI_ISL_706248\|20 | ···················· | ···················· | ···················· | ···················· | ···················· | ···················· |
| W | 3274 hCoV-19/England/MILK-B3DC1A/2020\|EPI_ISL_706249\|20 | ···················· | ···················· | ···················· | ···················· | ···················· | ···················· |
| W | 3275 hCoV-19/Thailand/Chonburi-SQ-NT0031/2020\|EPI_ISL_7 | ···················· | ···················· | ···················· | ···················· | ···················· | ···················· |
| W | 3276 hCoV-19/Thailand/Chonburi-SQ-TS0001/2020\|EPI_ISL_7 | ···················· | ···················· | ···················· | ···················· | ···················· | ···················· |
| W | 3277 hCoV-19/Thailand/Chonburi-SQ-NP0073/2020\|EPI_ISL_7 | ···················· | ···················· | ···················· | ···················· | ···················· | ···················· |
| W | 3278 hCoV-19/Thailand/Chonburi-SQ-LN0002/2020\|EPI_ISL_7 | ···················· | ···················· | ···················· | ···················· | ···················· | ···················· |
| W | 3279 hCoV-19/Thailand/Chonburi-SQ-RF0001/2020\|EPI_ISL_7 | ···················· | ···················· | ···················· | ···················· | ···················· | ···················· |
| W | 3280 hCoV-19/Thailand/Chonburi-SQ-SM0001/2020\|EPI_ISL_7 | ···················· | ···················· | ···················· | ···················· | ···················· | ···················· |
| W | 3281 hCoV-19/Thailand/Chonburi-SQ-TB0062/2020\|EPI_ISL_7 | ···················· | ···················· | ···················· | ···················· | ···················· | ···················· |
| W | 3282 hCoV-19/Thailand/phitsanulok-NT0076/2020\|EPI_ISL_7 | ···················· | ·A·················· | ···················· | ···················· | ···················· | ···A················ |
| W | 3283 hCoV-19/Thailand/phitsanulok-MT0115/2020\|EPI_ISL_7 | ···················· | ·A·················· | ···················· | ···················· | ···················· | ···A················ |
| W | 3284 hCoV-19/Thailand/Bangkok-PH7928/2020\|EPI_ISL_70881 | ···················· | ·A·················· | ···················· | ···················· | ···················· | ···A················ |
| W | 3285 hCoV-19/Thailand/Bangkok-Ge04/2020\|EPI_ISL_708813\| | ···················· | ···················· | ···················· | ···················· | ···················· | ···················· |
| W | 3286 hCoV-19/Thailand/Bangkok-14712/2020\|EPI_ISL_708815 | ···················· | ·A·················· | ···················· | ···················· | ···················· | ···A················ |
| W | 3287 hCoV-19/Thailand/Bangkok-14918/2020\|EPI_ISL_708816 | ···················· | ···················· | ···················· | ···················· | ···················· | ···················· |
| W | 3288 hCoV-19/Thailand/Bangkok-15401/2020\|EPI_ISL_708817 | ···················· | ···················· | ···················· | ···················· | ···················· | ···················· |
| W | 3289 hCoV-19/Thailand/Bangkok-14765/2020\|EPI_ISL_708818 | ···················· | ···················· | ···················· | ···················· | ···················· | ···················· |
| W | 3290 hCoV-19/Thailand/Bangkok-16110/2020\|EPI_ISL_708819 | ···················· | ···················· | ···················· | ···················· | ···················· | ···················· |
| W | 3291 hCoV-19/Thailand/Bangkok-Am01/2020\|EPI_ISL_708820\| | ···················· | ···················· | ···················· | ···················· | ···················· | ···················· |
| W | 3292 hCoV-19/Thailand/Bangkok-Ja08/2020\|EPI_ISL_708822\| | ···················· | ···················· | ···················· | ···················· | ···················· | ···················· |
| W | 3293 hCoV-19/Thailand/Bangkok-Tho07/2020\|EPI_ISL_708823 | ···················· | ···················· | ···················· | ···················· | ···················· | ···················· |
| W | 3294 hCoV-19/Thailand/Bangkok-Geo05/2020\|EPI_ISL_708824 | ···················· | ···················· | ···················· | ···················· | ···················· | ···················· |
| W | 3295 hCoV-19/Thailand/Bangkok-GA03/2020\|EPI_ISL_708826\| | ···················· | ···················· | ···················· | ···················· | ···················· | ···················· |
| W | 3296 hCoV-19/Pakistan/NIBD-01-KHI/2020\|EPI_ISL_708839\|2 | ···················· | ·A·················· | ···················· | ···················· | ···················· | ···A················ |
| W | 3297 hCoV-19/Pakistan/NIBD-02-KHI/2020\|EPI_ISL_708840\|2 | ···················· | ·A·················· | ···················· | ···················· | ···················· | ···A················ |
| W | 3298 hCoV-19/England/ALDP-BE9011/2020\|EPI_ISL_708845\|20 | ···················· | ···················· | ···················· | ···················· | ···················· | ···················· |
| W | 3299 hCoV-19/England/ALDP-BE955E/2020\|EPI_ISL_708846\|20 | ···················· | ···················· | ···················· | ···················· | ···················· | ···················· |
| W | 3300 hCoV-19/England/MILK-BEE252/2020\|EPI_ISL_708847\|20 | ···················· | ···················· | ···················· | ···················· | ···················· | ···················· |
| W | 3301 hCoV-19/England/MILK-BEE4B6/2020\|EPI_ISL_708848\|20 | ···················· | ···················· | ···················· | ···················· | ···················· | ···················· |

| | | | | | | | |
|---|---|---|---|---|---|---|---|
| W | 3302 hCoV-19/England/MILK-BEE29E/2020\|EPI_ISL_708850\|20 | ···················· | ···················· | ···················· | ···················· | ···················· | ···················· |
| W | 3303 hCoV-19/England/ALDP-BE8ECD/2020\|EPI_ISL_708851\|20 | ···················· | ···················· | ···················· | ···················· | ···················· | ···················· |
| W | 3304 hCoV-19/England/ALDP-BE8E45/2020\|EPI_ISL_708852\|20 | ···················· | ···················· | ···················· | ···················· | ···················· | ···················· |
| W | 3305 hCoV-19/England/ALDP-BE8EEB/2020\|EPI_ISL_708853\|20 | ···················· | ···················· | ···················· | ···················· | ···················· | ···················· |
| W | 3306 hCoV-19/England/ALDP-BE8FD9/2020\|EPI_ISL_708854\|20 | ···················· | ···················· | ···················· | ···················· | ···················· | ···················· |
| W | 3307 hCoV-19/England/QEUH-BEAFE6/2020\|EPI_ISL_708855\|20 | ···················· | ·A·················· | ···················· | ···················· | ···················· | ···················· |
| W | 3308 hCoV-19/England/MILK-BEE4F2/2020\|EPI_ISL_708857\|20 | ···················· | ···················· | ···················· | ···················· | ···················· | ···················· |
| W | 3309 hCoV-19/England/ALDP-BE9819/2020\|EPI_ISL_708861\|20 | ···················· | ···················· | ···················· | ···················· | ···················· | ···················· |
| W | 3310 hCoV-19/England/MILK-BEE2BC/2020\|EPI_ISL_708862\|20 | ···················· | ···················· | ···················· | ···················· | ···················· | ···················· |
| W | 3311 hCoV-19/England/ALDP-BE9002/2020\|EPI_ISL_708864\|20 | ···················· | ···················· | ···················· | ···················· | ···················· | ···················· |
| W | 3312 hCoV-19/England/ALDP-BE8FF7/2020\|EPI_ISL_708865\|20 | ···················· | ···················· | ···················· | ···················· | ···················· | ···················· |
| W | 3313 hCoV-19/England/ALDP-BE8E09/2020\|EPI_ISL_708866\|20 | ···················· | ···················· | ···················· | ···················· | ···················· | ···················· |
| W | 3314 hCoV-19/England/ALDP-BE904E/2020\|EPI_ISL_708867\|20 | ···················· | ···················· | ···················· | ···················· | ···················· | ···················· |
| W | 3315 hCoV-19/England/ALDP-BE912D/2020\|EPI_ISL_708868\|20 | ···················· | ···················· | ···················· | ···················· | ···················· | ···················· |
| W | 3316 hCoV-19/England/ALDP-BE9530/2020\|EPI_ISL_708870\|20 | ···················· | ···················· | ···················· | ···················· | ···················· | ···················· |
| W | 3317 hCoV-19/England/MILK-BEE2AD/2020\|EPI_ISL_708871\|20 | ···················· | ···················· | ···················· | ···················· | ···················· | ···················· |
| W | 3318 hCoV-19/England/ALDP-BE91B4/2020\|EPI_ISL_708872\|20 | ···················· | ···················· | ···················· | ···················· | ···················· | ···················· |
| W | 3319 hCoV-19/England/ALDP-BE922A/2020\|EPI_ISL_708877\|20 | ···················· | ···················· | ···················· | ···················· | ···················· | ···················· |
| W | 3320 hCoV-19/England/QEUH-BEB45F/2020\|EPI_ISL_708890\|20 | ···················· | ···················· | ···················· | ···················· | ···················· | ···················· |
| W | 3321 hCoV-19/England/ALDP-BE97EF/2020\|EPI_ISL_708891\|20 | ···················· | ···················· | ···················· | ···················· | ···················· | ···················· |
| W | 3322 hCoV-19/England/ALDP-BE8F51/2020\|EPI_ISL_708892\|20 | ···················· | ···················· | ···················· | ···················· | ···················· | ···················· |
| W | 3323 hCoV-19/England/ALDP-BE911E/2020\|EPI_ISL_708894\|20 | ···················· | ···················· | ···················· | ···················· | ···················· | ···················· |
| W | 3324 hCoV-19/England/ALDP-BE954F/2020\|EPI_ISL_708895\|20 | ···················· | ···················· | ···················· | ···················· | ···················· | ···················· |
| W | 3325 hCoV-19/England/MILK-BEE50E/2020\|EPI_ISL_708896\|20 | ···················· | ···················· | ···················· | ···················· | ···················· | ···················· |
| W | 3326 hCoV-19/England/MILK-BEE4E3/2020\|EPI_ISL_708897\|20 | ···················· | ···················· | ···················· | ···················· | ···················· | ···················· |
| W | 3327 hCoV-19/England/MILK-BEE35F/2020\|EPI_ISL_708898\|20 | ···················· | ···················· | ···················· | ···················· | ······T············· | ···················· |
| W | 3328 hCoV-19/England/ALDP-BE8FAC/2020\|EPI_ISL_708899\|20 | ···················· | ···················· | ···················· | ···················· | ···················· | ···················· |
| W | 3329 hCoV-19/USA/AZ-TG491556/2020\|EPI_ISL_695169\|2020-0 | ···················· | ···················· | ···················· | ···················· | ···················· | ···················· |
| W | 3330 hCoV-19/USA/AZ-TG491557/2020\|EPI_ISL_695170\|2020-0 | ···················· | ···················· | ···················· | ···················· | ···················· | ···················· |
| W | 3331 hCoV-19/USA/AZ-TG491559/2020\|EPI_ISL_695172\|2020-0 | ···················· | ···················· | ···················· | ···················· | ···················· | ···················· |
| W | 3332 hCoV-19/USA/AZ-TG491560/2020\|EPI_ISL_695173\|2020-0 | ···················· | ·A·················· | ···················· | ···················· | ···················· | ···················· |
| W | 3333 hCoV-19/USA/AZ-TG491578/2020\|EPI_ISL_695177\|2020-0 | ···················· | ··C················· | ···················· | ···················· | ···················· | ···················· |
| W | 3334 hCoV-19/USA/AZ-TG491600/2020\|EPI_ISL_695186\|2020-0 | ···················· | ···················· | ···················· | ···················· | ···················· | ···················· |
| W | 3335 hCoV-19/USA/AZ-TG498783/2020\|EPI_ISL_695271\|2020-0 | ···················· | ·A·················· | ···················· | ···················· | ···················· | ···················· |
| W | 3336 hCoV-19/USA/AZ-TG498910/2020\|EPI_ISL_695272\|2020-0 | ···················· | ·A·················· | ···················· | ···················· | ···················· | ···················· |
| W | 3337 hCoV-19/USA/AZ-TG498998/2020\|EPI_ISL_695273\|2020-0 | ···················· | ·A·················· | ···················· | ···················· | ···················· | ···················· |
| W | 3338 hCoV-19/USA/AZ-TG502386/2020\|EPI_ISL_695282\|2020-0 | ···················· | ·A·················· | ···················· | ···················· | ···················· | ···················· |
| W | 3339 hCoV-19/USA/AZ-TG502397/2020\|EPI_ISL_695283\|2020-0 | ···················· | ···················· | ···················· | ···················· | ···················· | ···················· |
| W | 3340 hCoV-19/USA/AZ-TG502522/2020\|EPI_ISL_695284\|2020-0 | ···················· | ···················· | ···················· | ···················· | ···················· | ···················· |
| W | 3341 hCoV-19/USA/AZ-TG502713/2020\|EPI_ISL_695285\|2020-0 | ···················· | ·A·················· | ···················· | ···················· | ···················· | ···················· |
| W | 3346 hCoV-19/South Africa/NHLS-UCT-GS-7041-KRISP/2020\|E | ···················· | ···················· | ···················· | ···················· | ···················· | ···················· |
| W | 3347 hCoV-19/South Africa/NHLS-UCT-GS-7030-KRISP/2020\|E | ···················· | ·A·················· | ···················· | ···················· | ···················· | ···················· |
| W | 3352 hCoV-19/Ecuador/USFQ-539/2020\|EPI_ISL_697800\|2020- | ···················· | ···················· | ···················· | ···················· | ···················· | ···················· |
| W | 3353 hCoV-19/India/MH-ACTREC-452/2020\|EPI_ISL_700100\|20 | ···················· | ···················· | ···················· | ···················· | ···················· | ···················· |
| W | 3354 hCoV-19/India/MH-ACTREC-453/2020\|EPI_ISL_700101\|20 | ···················· | ···················· | ···················· | ···················· | ···················· | ···················· |
| W | 3355 hCoV-19/India/MH-ACTREC-454/2020\|EPI_ISL_700102\|20 | ···················· | ···················· | ···················· | ···················· | ···················· | ···················· |
| W | 3356 hCoV-19/India/MH-ACTREC-455/2020\|EPI_ISL_700103\|20 | ···················· | ···················· | ···················· | ···················· | ···················· | ···················· |
| W | 3357 hCoV-19/India/MH-ACTREC-456/2020\|EPI_ISL_700104\|20 | ···················· | ···················· | ···················· | ···················· | ···················· | ···················· |
| W | 3358 hCoV-19/India/MH-ACTREC-457/2020\|EPI_ISL_700105\|20 | ···················· | ···················· | ···················· | ···················· | ···················· | ···················· |
| W | 3359 hCoV-19/India/MH-ACTREC-458/2020\|EPI_ISL_700106\|20 | ···················· | ···················· | ···················· | ···················· | ···················· | ···················· |
| W | 3360 hCoV-19/India/MH-ACTREC-459/2020\|EPI_ISL_700107\|20 | ···················· | ·A·················· | ···················· | ···················· | ···················· | ···················· |
| W | 3361 hCoV-19/India/MH-ACTREC-460/2020\|EPI_ISL_700108\|20 | ···················· | ···················· | ···················· | ···················· | ···················· | ···················· |
| W | 3364 hCoV-19/India/MH-ACTREC-463/2020\|EPI_ISL_700111\|20 | ···················· | ···················· | ···················· | ···················· | ···················· | ···················· |
| W | 3365 hCoV-19/India/MH-ACTREC-464/2020\|EPI_ISL_700112\|20 | ···················· | ···················· | ···················· | ···················· | ···················· | ···················· |
| W | 3366 hCoV-19/India/MH-ACTREC-465/2020\|EPI_ISL_700113\|20 | ···················· | ···················· | ···················· | ···················· | ···················· | ···················· |
| W | 3367 hCoV-19/India/MH-ACTREC-466/2020\|EPI_ISL_700114\|20 | ···················· | ···················· | ···················· | ···················· | ···················· | ···················· |
| W | 3368 hCoV-19/India/MH-ACTREC-467/2020\|EPI_ISL_700115\|20 | ···················· | ···················· | ···················· | ···················· | ···················· | ···················· |
| W | 3369 hCoV-19/India/MH-ACTREC-468/2020\|EPI_ISL_700116\|20 | ···················· | ···················· | ···················· | ···················· | ···················· | ···················· |
| W | 3370 hCoV-19/India/MH-ACTREC-469/2020\|EPI_ISL_700117\|20 | ···················· | ···················· | ···················· | ···················· | ···················· | ···················· |
| W | 3371 hCoV-19/India/MH-ACTREC-470/2020\|EPI_ISL_700118\|20 | ···················· | ···················· | ···················· | ···················· | ···················· | ···················· |
| W | 3372 hCoV-19/India/MH-ACTREC-471/2020\|EPI_ISL_700119\|20 | ···················· | ·A·················· | ···················· | ···················· | ···················· | ···················· |
| W | 3373 hCoV-19/India/MH-ACTREC-472/2020\|EPI_ISL_700120\|20 | ···················· | ·A·················· | ···················· | ···················· | ···················· | ···················· |
| W | 3374 hCoV-19/India/MH-ACTREC-473/2020\|EPI_ISL_700121\|20 | ···················· | ·A·················· | ···················· | ···················· | ···················· | ···················· |
| W | 3376 hCoV-19/India/MH-ACTREC-475/2020\|EPI_ISL_700123\|20 | ···················· | ·A·················· | ···················· | ···················· | ···················· | ···················· |
| W | 3377 hCoV-19/India/MH-ACTREC-476/2020\|EPI_ISL_700124\|20 | ···················· | ···················· | ···················· | ···················· | ···················· | ···················· |
| W | 3378 hCoV-19/India/MH-ACTREC-477/2020\|EPI_ISL_700125\|20 | ···················· | ···················· | ···················· | ···················· | ···················· | ···················· |
| W | 3379 hCoV-19/India/MH-ACTREC-478/2020\|EPI_ISL_700126\|20 | ···················· | ···················· | ···················· | ···················· | ···················· | ···················· |
| W | 3381 hCoV-19/India/MH-ACTREC-480/2020\|EPI_ISL_700128\|20 | ···················· | ···················· | ···················· | ···················· | ···················· | ···················· |
| W | 3382 hCoV-19/India/MH-ACTREC-481/2020\|EPI_ISL_700129\|20 | ···················· | ·A·················· | ···················· | ···················· | ···················· | ···················· |
| W | 3383 hCoV-19/India/MH-ACTREC-483/2020\|EPI_ISL_700131\|20 | ···················· | ·A·················· | ···················· | ···················· | ···················· | ···················· |
| W | 3385 hCoV-19/India/MH-ACTREC-485/2020\|EPI_ISL_700133\|20 | ···················· | ···················· | ···················· | ···················· | ···················· | ···················· |
| W | 3386 hCoV-19/India/MH-ACTREC-486/2020\|EPI_ISL_700134\|20 | ···················· | ···················· | ···················· | ···················· | ···················· | ···················· |
| W | 3387 hCoV-19/India/MH-ACTREC-487/2020\|EPI_ISL_700135\|20 | ···················· | ···················· | ···················· | ···················· | ···················· | ···················· |
| W | 3388 hCoV-19/India/MH-ACTREC-488/2020\|EPI_ISL_700136\|20 | ···················· | ···················· | ···················· | ···················· | ···················· | ···················· |
| W | 3389 hCoV-19/India/MH-ACTREC-489/2020\|EPI_ISL_700137\|20 | ···················· | ···················· | ···················· | ···················· | ···················· | ···················· |
| W | 3390 hCoV-19/India/MH-ACTREC-490/2020\|EPI_ISL_700138\|20 | ···················· | ·A·················· | ···················· | ···················· | ···················· | ···A················ |
| W | 3391 hCoV-19/India/MH-ACTREC-491/2020\|EPI_ISL_700139\|20 | ···················· | ···················· | ···················· | ···················· | ···················· | ···················· |
| W | 3392 hCoV-19/India/MH-ACTREC-492/2020\|EPI_ISL_700140\|20 | ···················· | ···················· | ···················· | ···················· | ···················· | ···················· |
| W | 3393 hCoV-19/India/MH-ACTREC-493/2020\|EPI_ISL_700141\|20 | ···················· | ·A·················· | ···················· | ···················· | ···················· | ···················· |
| W | 3394 hCoV-19/India/MH-ACTREC-494/2020\|EPI_ISL_700142\|20 | ···················· | ···················· | ···················· | ···················· | ···················· | ···················· |

| | | | | | | | | |
|---|---|---|---|---|---|---|---|---|
| W | 3395 | hCoV-19/India/MH-ACTREC-495/2020\|EPI_ISL_700143\|20 | ···················· | ···················· | ····················· | ···················· | ···················· | ···················· |
| W | 3396 | hCoV-19/India/MH-ACTREC-496/2020\|EPI_ISL_700144\|20 | ···················· | ···················· | ····················· | ···················· | ···················· | ···················· |
| W | 3397 | hCoV-19/India/MH-ACTREC-497/2020\|EPI_ISL_700145\|20 | ···················· | ···················· | ····················· | ···················· | ···················· | ···················· |
| W | 3398 | hCoV-19/India/MH-ACTREC-498/2020\|EPI_ISL_700146\|20 | ···················· | ···················· | ····················· | ···················· | ···················· | ···················· |
| W | 3399 | hCoV-19/India/MH-ACTREC-499/2020\|EPI_ISL_700147\|20 | ···················· | ···················· | ····················· | ···················· | ···················· | ···················· |
| W | 3400 | hCoV-19/India/MH-ACTREC-500/2020\|EPI_ISL_700148\|20 | ···················· | ···················· | ····················· | ···················· | ···················· | ···················· |
| W | 3401 | hCoV-19/India/MH-ACTREC-501/2020\|EPI_ISL_700149\|20 | ···················· | ···················· | ····················· | ···················· | ···················· | ···················· |
| W | 3402 | hCoV-19/India/MH-ACTREC-502/2020\|EPI_ISL_700150\|20 | ···················· | ···················· | ····················· | ···················· | ···················· | ···················· |
| W | 3403 | hCoV-19/India/MH-ACTREC-503/2020\|EPI_ISL_700151\|20 | ···················· | ···················· | ····················· | ···················· | ···················· | ···················· |
| W | 3404 | hCoV-19/India/MH-ACTREC-504/2020\|EPI_ISL_700152\|20 | ···················· | ···················· | ····················· | ···················· | ···················· | ···················· |
| W | 3405 | hCoV-19/India/MH-ACTREC-505/2020\|EPI_ISL_700153\|20 | ···················· | ···················· | ····················· | ···················· | ···················· | ···················· |
| W | 3406 | hCoV-19/India/MH-ACTREC-506/2020\|EPI_ISL_700154\|20 | ···················· | ···················· | ····················· | ···················· | ···················· | ···················· |
| W | 3407 | hCoV-19/India/MH-ACTREC-507/2020\|EPI_ISL_700155\|20 | ···················· | ···················· | ····················· | ···················· | ···················· | ···················· |
| W | 3408 | hCoV-19/India/MH-ACTREC-508/2020\|EPI_ISL_700156\|20 | ···················· | ···················· | ····················· | ···················· | ···················· | ···················· |
| W | 3409 | hCoV-19/India/MH-ACTREC-509/2020\|EPI_ISL_700157\|20 | ···················· | ···················· | ····················· | ···················· | ···················· | ···················· |
| W | 3410 | hCoV-19/India/MH-ACTREC-510/2020\|EPI_ISL_700158\|20 | ···················· | ···················· | ····················· | ···················· | ···················· | ···················· |
| W | 3411 | hCoV-19/India/MH-ACTREC-513/2020\|EPI_ISL_700160\|20 | ···················· | ···················· | ····················· | ···················· | ···················· | ···················· |
| W | 3412 | hCoV-19/India/MH-ACTREC-514/2020\|EPI_ISL_700161\|20 | ···················· | ···················· | ····················· | ···················· | ···················· | ···················· |
| W | 3413 | hCoV-19/India/MH-ACTREC-515/2020\|EPI_ISL_700162\|20 | ···················· | ···················· | ····················· | ···················· | ···················· | ···················· |
| W | 3414 | hCoV-19/India/MH-ACTREC-516/2020\|EPI_ISL_700163\|20 | ···················· | ···················· | ····················· | ···················· | ···················· | ···················· |
| W | 3415 | hCoV-19/India/MH-ACTREC-518/2020\|EPI_ISL_700164\|20 | ···················· | ···················· | ····················· | ···················· | ···················· | ···················· |
| W | 3416 | hCoV-19/India/MH-ACTREC-519/2020\|EPI_ISL_700165\|20 | ···················· | ···················· | ····················· | ···················· | ···················· | ···················· |
| W | 3417 | hCoV-19/India/MH-ACTREC-520/2020\|EPI_ISL_700166\|20 | ···················· | ···················· | ····················· | ···················· | ···················· | ···················· |
| W | 3418 | hCoV-19/India/MH-ACTREC-521/2020\|EPI_ISL_700167\|20 | ···················· | ···················· | ····················· | ···················· | ···················· | ···················· |
| W | 3419 | hCoV-19/India/MH-ACTREC-522/2020\|EPI_ISL_700168\|20 | ···················· | ···················· | ····················· | ···················· | ···················· | ···················· |
| W | 3420 | hCoV-19/India/MH-ACTREC-523/2020\|EPI_ISL_700169\|20 | ···················· | ···················· | ····················· | ···················· | ···················· | ···················· |
| W | 3421 | hCoV-19/India/MH-ACTREC-524/2020\|EPI_ISL_700170\|20 | ···················· | ···················· | ····················· | ···················· | ···················· | ···················· |
| W | 3422 | hCoV-19/India/MH-ACTREC-525/2020\|EPI_ISL_700171\|20 | ···················· | ···················· | ····················· | ···················· | ···················· | ···················· |
| W | 3423 | hCoV-19/India/MH-ACTREC-526/2020\|EPI_ISL_700172\|20 | ···················· | ···················· | ····················· | ···················· | ···················· | ···················· |
| W | 3424 | hCoV-19/India/MH-ACTREC-527/2020\|EPI_ISL_700173\|20 | ···················· | ···················· | ····················· | ···················· | ···················· | ···················· |
| W | 3425 | hCoV-19/India/MH-ACTREC-529/2020\|EPI_ISL_700175\|20 | ···················· | ···················· | ····················· | ···················· | ···················· | ···················· |
| W | 3426 | hCoV-19/India/MH-ACTREC-530/2020\|EPI_ISL_700176\|20 | ···················· | ···················· | ····················· | ···················· | ···················· | ···················· |
| W | 3427 | hCoV-19/India/MH-ACTREC-531/2020\|EPI_ISL_700177\|20 | ···················· | ···················· | ····················· | ···················· | ···················· | ···················· |
| W | 3428 | hCoV-19/India/MH-ACTREC-532/2020\|EPI_ISL_700178\|20 | ···················· | ···················· | ····················· | ···················· | ···················· | ···················· |
| W | 3429 | hCoV-19/India/MH-ACTREC-533/2020\|EPI_ISL_700179\|20 | ···················· | ···················· | ····················· | ···················· | ···················· | ···················· |
| W | 3431 | hCoV-19/India/MH-ACTREC-535/2020\|EPI_ISL_700181\|20 | ···················· | ···················· | ····················· | ···················· | ···················· | ···················· |
| W | 3432 | hCoV-19/India/MH-ACTREC-536/2020\|EPI_ISL_700182\|20 | ···················· | ···················· | ····················· | ···················· | ···················· | ···················· |
| W | 3433 | hCoV-19/India/MH-ACTREC-537/2020\|EPI_ISL_700183\|20 | ···················· | ·A·················· | ····················· | ···················· | ···················· | ···················· |
| W | 3434 | hCoV-19/India/MH-ACTREC-538/2020\|EPI_ISL_700184\|20 | ········T··········· | ···················· | ····················· | ···················· | ···················· | ···················· |
| W | 3435 | hCoV-19/India/MH-ACTREC-539/2020\|EPI_ISL_700185\|20 | ···················· | ···················· | ····················· | ···················· | ···················· | ···················· |
| W | 3436 | hCoV-19/India/MH-ACTREC-540/2020\|EPI_ISL_700186\|20 | ···················· | ···················· | ····················· | ···················· | ···················· | ···················· |
| W | 3437 | hCoV-19/India/MH-ACTREC-541/2020\|EPI_ISL_700187\|20 | ···················· | ···················· | ····················· | ···················· | ···················· | ···················· |
| W | 3438 | hCoV-19/India/MH-ACTREC-542/2020\|EPI_ISL_700188\|20 | ···················· | ···················· | ····················· | ···················· | ···················· | ···················· |
| W | 3439 | hCoV-19/India/MH-ACTREC-543/2020\|EPI_ISL_700189\|20 | ···················· | ···················· | ····················· | ···················· | ···················· | ···················· |
| W | 3440 | hCoV-19/India/MH-ACTREC-544/2020\|EPI_ISL_700190\|20 | ···················· | ···················· | ····················· | ···················· | ···················· | ···················· |
| W | 3441 | hCoV-19/India/MH-ACTREC-545/2020\|EPI_ISL_700191\|20 | ···················· | ···················· | ····················· | ···················· | ···················· | ···················· |
| W | 3442 | hCoV-19/India/MH-ACTREC-547/2020\|EPI_ISL_700192\|20 | ···················· | ···················· | ····················· | ···················· | ···················· | ···················· |
| W | 3443 | hCoV-19/India/MH-ACTREC-548/2020\|EPI_ISL_700193\|20 | ···················· | ···················· | ····················· | ···················· | ···················· | ···················· |
| W | 3444 | hCoV-19/India/MH-ACTREC-549/2020\|EPI_ISL_700194\|20 | ···················· | ···················· | ····················· | ···················· | ···················· | ···················· |
| W | 3445 | hCoV-19/India/MH-ACTREC-550/2020\|EPI_ISL_700195\|20 | ···················· | ·A·················· | ····················· | ···················· | ···················· | ···A················ |
| W | 3446 | hCoV-19/India/MH-ACTREC-551/2020\|EPI_ISL_700196\|20 | ···················· | ···················· | ····················· | ···················· | ···················· | ···················· |
| W | 3447 | hCoV-19/India/MH-ACTREC-552/2020\|EPI_ISL_700197\|20 | ···················· | ···················· | ····················· | ···················· | ···················· | ···················· |
| W | 3448 | hCoV-19/India/MH-ACTREC-553/2020\|EPI_ISL_700198\|20 | ···················· | ·A·················· | ····················· | ···················· | ···················· | ···················· |
| W | 3450 | hCoV-19/England/ALDP-BB32FC/2020\|EPI_ISL_702701\|20 | ···················· | ···················· | ····················· | ···················· | ···················· | ···················· |
| W | 3451 | hCoV-19/England/ALDP-BB5DF9/2020\|EPI_ISL_702702\|20 | ···················· | ···················· | ····················· | ···················· | ···················· | ···················· |
| W | 3452 | hCoV-19/England/ALDP-BB2891/2020\|EPI_ISL_702704\|20 | ···················· | ···················· | ····················· | ···················· | ···················· | ···················· |
| W | 3453 | hCoV-19/England/ALDP-BB3247/2020\|EPI_ISL_702706\|20 | ···················· | ···················· | ····················· | ···················· | ······T············· | ···················· |
| W | 3454 | hCoV-19/England/ALDP-BB5913/2020\|EPI_ISL_702709\|20 | ···················· | ···················· | ····················· | ···················· | ···················· | ···················· |
| W | 3455 | hCoV-19/England/ALDP-BB2DFC/2020\|EPI_ISL_702711\|20 | ···················· | ···················· | ····················· | ···················· | ······T············· | ···················· |
| W | 3456 | hCoV-19/England/ALDP-BB27A3/2020\|EPI_ISL_702714\|20 | ···················· | ···················· | ····················· | ···················· | ···················· | ···················· |
| W | 3457 | hCoV-19/England/ALDP-BB9478/2020\|EPI_ISL_702715\|20 | ···················· | ···················· | ····················· | ···················· | ···················· | ···················· |
| W | 3458 | hCoV-19/England/ALDP-BB8BB0/2020\|EPI_ISL_702717\|20 | ···················· | ···················· | ····················· | ···················· | ···················· | ···················· |
| W | 3459 | hCoV-19/England/ALDP-BB34F6/2020\|EPI_ISL_702719\|20 | ···················· | ···················· | ····················· | ···················· | ···················· | ···················· |
| W | 3460 | hCoV-19/England/ALDP-BB5D44/2020\|EPI_ISL_702720\|20 | ···················· | ···················· | ····················· | ···················· | ···················· | ···················· |
| W | 3461 | hCoV-19/England/ALDP-BB2E35/2020\|EPI_ISL_702722\|20 | ···················· | ···················· | ····················· | ···················· | ···················· | ···················· |
| W | 3462 | hCoV-19/England/ALDP-BB9A94/2020\|EPI_ISL_702724\|20 | ···················· | ·A·················· | ····················· | ···················· | ···················· | ···················· |
| W | 3463 | hCoV-19/England/ALDP-BB28DD/2020\|EPI_ISL_702725\|20 | ···················· | ···················· | ····················· | ···················· | ···················· | ···················· |
| W | 3464 | hCoV-19/England/ALDP-BB9D5F/2020\|EPI_ISL_702727\|20 | ···················· | ···················· | ····················· | ···················· | ···················· | ···················· |
| W | 3465 | hCoV-19/England/ALDP-BB9997/2020\|EPI_ISL_702730\|20 | ···················· | ···················· | ····················· | ···················· | ···················· | ···················· |
| W | 3466 | hCoV-19/England/ALDP-BB39AC/2020\|EPI_ISL_702733\|20 | ···················· | ···················· | ····················· | ···················· | ···················· | ···················· |
| W | 3467 | hCoV-19/England/ALDP-BB8C35/2020\|EPI_ISL_702735\|20 | ···················· | ···················· | ····················· | ···················· | ···················· | ···················· |
| W | 3468 | hCoV-19/England/ALDP-BB8BCF/2020\|EPI_ISL_702737\|20 | ···················· | ···················· | ····················· | ···················· | ···················· | ···················· |
| W | 3469 | hCoV-19/England/ALDP-BB34AB/2020\|EPI_ISL_702740\|20 | ···················· | ···················· | ····················· | ···················· | ···················· | ···················· |
| W | 3470 | hCoV-19/England/ALDP-BB357B/2020\|EPI_ISL_702741\|20 | ···················· | ···················· | ····················· | ···················· | ···················· | ···················· |
| W | 3471 | hCoV-19/England/ALDP-BB8BED/2020\|EPI_ISL_702743\|20 | ···················· | ···················· | ····················· | ···················· | ···················· | ···················· |
| W | 3472 | hCoV-19/England/ALDP-BB39BB/2020\|EPI_ISL_702745\|20 | ···················· | ·A·················· | ····················· | ···················· | ···················· | ···················· |
| W | 3473 | hCoV-19/England/ALDP-BB97E8/2020\|EPI_ISL_702748\|20 | ···················· | ···················· | ····················· | ···················· | ···················· | ···················· |
| W | 3474 | hCoV-19/England/ALDP-BB8E5D/2020\|EPI_ISL_702750\|20 | ···················· | ···················· | ····················· | ···················· | ······T············· | ···················· |
| W | 3475 | hCoV-19/England/ALDP-BB361E/2020\|EPI_ISL_702752\|20 | ···················· | ···················· | ····················· | ···················· | ···················· | ···················· |
| W | 3476 | hCoV-19/England/ALDP-BB31C2/2020\|EPI_ISL_702755\|20 | ···················· | ·A·················· | ····················· | ···················· | ···················· | ···A················ |

| | | | | | | | | |
|---|---|---|---|---|---|---|---|---|
| W | 3477 | hCoV-19/England/ALDP-BB986D/2020\|EPI_ISL_702757\|20 | ···················· | ···················· | ····················· | ···················· | ···················· | ···················· |
| W | 3478 | hCoV-19/England/ALDP-BB59C8/2020\|EPI_ISL_702759\|20 | ···················· | ···················· | ····················· | ···················· | ···················· | ···················· |
| W | 3479 | hCoV-19/England/ALDP-BB9C25/2020\|EPI_ISL_702760\|20 | ···················· | ···················· | ····················· | ···················· | ···················· | ···················· |
| W | 3480 | hCoV-19/England/ALDP-BB5579/2020\|EPI_ISL_702764\|20 | ···················· | ···················· | ····················· | ···················· | ···················· | ···················· |
| W | 3481 | hCoV-19/England/ALDP-BB5333/2020\|EPI_ISL_702765\|20 | ···················· | ···················· | ····················· | ···················· | ···················· | ···················· |
| W | 3482 | hCoV-19/England/ALDP-BB8C08/2020\|EPI_ISL_702767\|20 | ···················· | ···················· | ···········T········· | ···················· | ···················· | ···················· |
| W | 3483 | hCoV-19/England/ALDP-BB59AA/2020\|EPI_ISL_702768\|20 | ···················· | ···················· | ····················· | ···················· | ···················· | ···················· |
| W | 3484 | hCoV-19/England/ALDP-BB3265/2020\|EPI_ISL_702772\|20 | ···················· | ···················· | ····················· | ···················· | ······T············· | ···················· |
| W | 3485 | hCoV-19/England/ALDP-BB311D/2020\|EPI_ISL_702773\|20 | ···················· | ···················· | ····················· | ···················· | ···················· | ···················· |
| W | 3486 | hCoV-19/England/ALDP-BB2846/2020\|EPI_ISL_702775\|20 | ···················· | ···················· | ····················· | ···················· | ···················· | ···················· |
| W | 3487 | hCoV-19/England/ALDP-BB992E/2020\|EPI_ISL_702777\|20 | ···················· | ···················· | ····················· | ···················· | ···················· | ···················· |
| W | 3488 | hCoV-19/England/ALDP-BB2E26/2020\|EPI_ISL_702778\|20 | ···················· | ···················· | ····················· | ···················· | ···················· | ···················· |
| W | 3489 | hCoV-19/England/ALDP-BB9CDA/2020\|EPI_ISL_702780\|20 | ···················· | ···················· | ····················· | ···················· | ···················· | ···················· |
| W | 3490 | hCoV-19/England/ALDP-BB5773/2020\|EPI_ISL_702781\|20 | ···················· | ···················· | ····················· | ···················· | ···················· | ···················· |
| W | 3491 | hCoV-19/England/ALDP-BB27FE/2020\|EPI_ISL_702783\|20 | ···················· | ···················· | ····················· | ···················· | ···················· | ···················· |
| W | 3492 | hCoV-19/England/ALDP-BB3960/2020\|EPI_ISL_702785\|20 | ···················· | ···················· | ····················· | ···················· | ···················· | ···················· |
| W | 3493 | hCoV-19/England/ALDP-BB2837/2020\|EPI_ISL_702786\|20 | ···················· | ···················· | ····················· | ···················· | ···················· | ···················· |
| W | 3494 | hCoV-19/England/ALDP-BB98A9/2020\|EPI_ISL_702788\|20 | ···················· | ···················· | ····················· | ···················· | ···················· | ···················· |
| W | 3495 | hCoV-19/England/ALDP-BB310E/2020\|EPI_ISL_702796\|20 | ···················· | ···················· | ····················· | ···················· | ···················· | ···················· |
| W | 3496 | hCoV-19/England/ALDP-BBACE8/2020\|EPI_ISL_702797\|20 | ···················· | ···················· | ····················· | ···················· | ···················· | ···················· |
| W | 3497 | hCoV-19/England/ALDP-BB2828/2020\|EPI_ISL_702799\|20 | ···················· | ···················· | ····················· | ···················· | ···················· | ···A················ |
| W | 3498 | hCoV-19/England/MILK-B3CFF4/2020\|EPI_ISL_706212\|20 | ···················· | ···················· | ····················· | ···················· | ···················· | ···················· |
| W | 3499 | hCoV-19/England/MILK-B3D00F/2020\|EPI_ISL_706213\|20 | ···················· | ·A·················· | ····················· | ···················· | ···················· | ···A················ |
| W | 3500 | hCoV-19/England/MILK-B3D05A/2020\|EPI_ISL_706215\|20 | ···················· | ···················· | ····················· | ···················· | ···················· | ···················· |
| W | 3501 | hCoV-19/England/MILK-B3D069/2020\|EPI_ISL_706216\|20 | ···················· | ···················· | ····················· | ···················· | ···················· | ···················· |
| W | 3502 | hCoV-19/England/MILK-B3D096/2020\|EPI_ISL_706217\|20 | ···················· | ···················· | ····················· | ···················· | ···················· | ···················· |
| W | 3503 | hCoV-19/England/MILK-B3D0B4/2020\|EPI_ISL_706218\|20 | ···················· | ···················· | ····················· | ···················· | ···················· | ···················· |
| W | 3504 | hCoV-19/England/MILK-B3D0C3/2020\|EPI_ISL_706219\|20 | ···················· | ···················· | ····················· | ···················· | ···················· | ···················· |
| W | 3505 | hCoV-19/England/MILK-B3D0D2/2020\|EPI_ISL_706220\|20 | ···················· | ···················· | ····················· | ···················· | ···················· | ···················· |
| W | 3506 | hCoV-19/England/MILK-B3D10C/2020\|EPI_ISL_706221\|20 | ···················· | ···················· | ····················· | ···················· | ···················· | ···················· |
| W | 3507 | hCoV-19/England/MILK-B3D12A/2020\|EPI_ISL_706222\|20 | ···················· | ···················· | ····················· | ···················· | ···················· | ···················· |
| W | 3508 | hCoV-19/England/MILK-B3D148/2020\|EPI_ISL_706223\|20 | ···················· | ···················· | ····················· | ···················· | ···················· | ···················· |
| W | 3509 | hCoV-19/England/MILK-B3D157/2020\|EPI_ISL_706224\|20 | ···················· | ···················· | ····················· | ···················· | ···················· | ···················· |
| W | 3510 | hCoV-19/England/MILK-B3D166/2020\|EPI_ISL_706225\|20 | ···················· | ···················· | ····················· | ···················· | ···················· | ···················· |
| W | 3511 | hCoV-19/England/MILK-B3D1A2/2020\|EPI_ISL_706226\|20 | ···················· | ···················· | ····················· | ···················· | ···················· | ···················· |
| W | 3512 | hCoV-19/England/MILK-B3D2AF/2020\|EPI_ISL_706227\|20 | ···················· | ···················· | ····················· | ···················· | ···················· | ···················· |
| W | 3513 | hCoV-19/England/MILK-B3D597/2020\|EPI_ISL_706228\|20 | ···················· | ···················· | ····················· | ···················· | ···················· | ···················· |
| W | 3514 | hCoV-19/England/MILK-B3D5A6/2020\|EPI_ISL_706229\|20 | ···················· | ···················· | ····················· | ···················· | ···················· | ···················· |
| W | 3515 | hCoV-19/England/MILK-B3DC29/2020\|EPI_ISL_706250\|20 | ···················· | ···················· | ····················· | ···················· | ···················· | ···················· |
| W | 3516 | hCoV-19/England/MILK-B3DC65/2020\|EPI_ISL_706251\|20 | ···················· | ···················· | ····················· | ···················· | ···················· | ···················· |
| W | 3517 | hCoV-19/England/MILK-B3DC83/2020\|EPI_ISL_706252\|20 | ···················· | ···················· | ····················· | ···················· | ···················· | ···················· |
| W | 3518 | hCoV-19/England/MILK-B3DC92/2020\|EPI_ISL_706253\|20 | ···················· | ···················· | ····················· | ···················· | ···················· | ···················· |
| W | 3519 | hCoV-19/England/MILK-B3DCA1/2020\|EPI_ISL_706254\|20 | ···················· | ···················· | ····················· | ···················· | ···················· | ···················· |
| W | 3520 | hCoV-19/England/MILK-B3DCDE/2020\|EPI_ISL_706255\|20 | ···················· | ···················· | ····················· | ···················· | ···················· | ···················· |
| W | 3521 | hCoV-19/England/MILK-B3DD26/2020\|EPI_ISL_706256\|20 | ···················· | ···················· | ····················· | ···················· | ···················· | ···················· |
| W | 3522 | hCoV-19/England/MILK-B3DD44/2020\|EPI_ISL_706257\|20 | ···················· | ···················· | ····················· | ···················· | ···················· | ···················· |
| W | 3523 | hCoV-19/England/MILK-B3DD53/2020\|EPI_ISL_706258\|20 | ···················· | ···················· | ····················· | ···················· | ···················· | ···················· |
| W | 3524 | hCoV-19/England/MILK-B3DD62/2020\|EPI_ISL_706259\|20 | ···················· | ···················· | ····················· | ···················· | ···················· | ···················· |
| W | 3525 | hCoV-19/England/MILK-B3DD80/2020\|EPI_ISL_706260\|20 | ···················· | ···················· | ····················· | ···················· | ···················· | ···················· |
| W | 3526 | hCoV-19/England/MILK-B3E11A/2020\|EPI_ISL_706261\|20 | ···················· | ···················· | ····················· | ···················· | ···················· | ···················· |
| W | 3527 | hCoV-19/England/MILK-B3E147/2020\|EPI_ISL_706262\|20 | ···················· | ···················· | ····················· | ···················· | ···················· | ···················· |
| W | 3528 | hCoV-19/England/MILK-B3E165/2020\|EPI_ISL_706263\|20 | ···················· | ···················· | ····················· | ···················· | ···················· | ···················· |
| W | 3529 | hCoV-19/England/MILK-B3E174/2020\|EPI_ISL_706264\|20 | ···················· | ···················· | ····················· | ···················· | ···················· | ···················· |
| W | 3530 | hCoV-19/England/MILK-B3E1A1/2020\|EPI_ISL_706265\|20 | ···················· | ···················· | ····················· | ···················· | ···················· | ···················· |
| W | 3531 | hCoV-19/England/MILK-B3E226/2020\|EPI_ISL_706266\|20 | ···················· | ···················· | ····················· | ···················· | ···················· | ···················· |
| W | 3532 | hCoV-19/England/MILK-B3E235/2020\|EPI_ISL_706267\|20 | ···················· | ···················· | ····················· | ···················· | ···················· | ···················· |
| W | 3533 | hCoV-19/England/MILK-B3E253/2020\|EPI_ISL_706268\|20 | ···················· | ···················· | ····················· | ···················· | ···················· | ···················· |
| W | 3534 | hCoV-19/England/MILK-B3E323/2020\|EPI_ISL_706269\|20 | ···················· | ···················· | ····················· | ···················· | ···················· | ···················· |
| W | 3535 | hCoV-19/England/MILK-B3E341/2020\|EPI_ISL_706270\|20 | ···················· | ···················· | ····················· | ···················· | ···················· | ···················· |
| W | 3536 | hCoV-19/England/MILK-B3E36F/2020\|EPI_ISL_706271\|20 | ···················· | ···················· | ····················· | ···················· | ···················· | ···················· |
| W | 3537 | hCoV-19/England/MILK-B3E37E/2020\|EPI_ISL_706272\|20 | ···················· | ···················· | ····················· | ···················· | ···················· | ···················· |
| W | 3538 | hCoV-19/England/MILK-B3E3AB/2020\|EPI_ISL_706273\|20 | ···················· | ···················· | ····················· | ···················· | ······T············· | ···················· |
| W | 3539 | hCoV-19/England/MILK-B3E3C9/2020\|EPI_ISL_706274\|20 | ···················· | ···················· | ····················· | ···················· | ···················· | ···················· |
| W | 3540 | hCoV-19/England/MILK-B3E3D8/2020\|EPI_ISL_706275\|20 | ···················· | ···················· | ····················· | ···················· | ···················· | ···················· |
| W | 3541 | hCoV-19/England/NORT-1B61773/2020\|EPI_ISL_706276\|2 | ···················· | ···················· | ····················· | ···················· | ···················· | ···················· |
| W | 3542 | hCoV-19/England/NORT-1B617BF/2020\|EPI_ISL_706277\|2 | ···················· | ···················· | ····················· | ···················· | ···················· | ···················· |
| W | 3543 | hCoV-19/England/NORT-1B617CE/2020\|EPI_ISL_706278\|2 | ···················· | ···················· | ····················· | ···················· | ···················· | ···················· |
| W | 3544 | hCoV-19/England/NORT-1B62824/2020\|EPI_ISL_706279\|2 | ···················· | ···················· | ····················· | ···················· | ····T··············· | ···················· |
| W | 3545 | hCoV-19/England/NORT-1B62AD3/2020\|EPI_ISL_706280\|2 | ···················· | ···················· | ····················· | ···················· | ···················· | ···················· |
| W | 3546 | hCoV-19/England/NORT-1B62AE2/2020\|EPI_ISL_706281\|2 | ···················· | ···················· | ····················· | ···················· | ···················· | ···················· |
| W | 3547 | hCoV-19/England/NORT-1B62AF1/2020\|EPI_ISL_706282\|2 | ···················· | ···················· | ····················· | ···················· | ···················· | ···················· |
| W | 3548 | hCoV-19/England/NORT-1B62B2B/2020\|EPI_ISL_706283\|2 | ···················· | ···················· | ····················· | ···················· | ···················· | ···················· |
| W | 3549 | hCoV-19/England/NORT-1B62B94/2020\|EPI_ISL_706284\|2 | ···················· | ···················· | ····················· | ···················· | ···················· | ···················· |
| W | 3550 | hCoV-19/England/NORT-1B62BA3/2020\|EPI_ISL_706285\|2 | ···················· | ···················· | ····················· | ···················· | ···················· | ···················· |
| W | 3551 | hCoV-19/England/NORT-1B70665/2020\|EPI_ISL_706286\|2 | ···················· | ···················· | ····················· | ···················· | ···················· | ···················· |
| W | 3552 | hCoV-19/England/NORT-1B70683/2020\|EPI_ISL_706288\|2 | ···················· | ···················· | ····················· | ···················· | ···················· | ···················· |
| W | 3553 | hCoV-19/England/NORT-1B706B0/2020\|EPI_ISL_706289\|2 | ···················· | ···················· | ····················· | ···················· | ···················· | ···················· |
| W | 3554 | hCoV-19/England/NORT-1B70744/2020\|EPI_ISL_706290\|2 | ···················· | ···················· | ····················· | ···················· | ···················· | ···················· |
| W | 3555 | hCoV-19/England/NORT-1B70753/2020\|EPI_ISL_706291\|2 | ···················· | ···················· | ····················· | ···················· | ···················· | ···················· |
| W | 3556 | hCoV-19/England/NORT-1B70902/2020\|EPI_ISL_706292\|2 | ···················· | ···················· | ····················· | ···················· | ···················· | ···················· |

W 3557 hCoV-19/England/NORT-1B7093F/2020|EPI_ISL_706293|2 ···················· ···················· ····················· ···················· ···················· ····················
W 3558 hCoV-19/England/NORT-1B709B7/2020|EPI_ISL_706294|2 ···················· ···················· ····················· ···················· ···················· ····················
W 3559 hCoV-19/England/NORT-1B70A4B/2020|EPI_ISL_706295|2 ···················· ···················· ····················· ···················· ···················· ····················
W 3560 hCoV-19/England/NORT-1B70A69/2020|EPI_ISL_706296|2 ···················· ···················· ····················· ···················· ···················· ····················
W 3561 hCoV-19/England/NORT-1B70EC7/2020|EPI_ISL_706297|2 ···················· ···················· ····················· ···················· ···················· ····················
W 3562 hCoV-19/England/NORT-1B70EE5/2020|EPI_ISL_706298|2 ···················· ···················· ····················· ···················· ···················· ····················
W 3563 hCoV-19/England/NORT-1B70EF4/2020|EPI_ISL_706299|2 ···················· ···················· ····················· ···················· ···················· ····················
W 3564 hCoV-19/England/ALDP-BE913C/2020|EPI_ISL_708876|20 ···················· ···················· ····················· ···················· ···················· ····················
W 3565 hCoV-19/England/ALDP-BE958B/2020|EPI_ISL_708878|20 ···················· ···················· ····················· ···················· ···················· ····················
W 3566 hCoV-19/England/MILK-BEE234/2020|EPI_ISL_708879|20 ···················· ···················· ····················· ···················· ···················· ····················
W 3567 hCoV-19/England/ALDP-BE963D/2020|EPI_ISL_708882|20 ···················· ···················· ····················· ···················· ···················· ····················
W 3568 hCoV-19/England/QEUH-BEB02E/2020|EPI_ISL_708884|20 ···················· ···················· ····················· ···················· ···················· ····················
W 3569 hCoV-19/England/ALDP-BE8EBE/2020|EPI_ISL_708886|20 ···················· ···················· ····················· ···················· ···················· ····················
W 3570 hCoV-19/England/ALDP-BE9503/2020|EPI_ISL_708887|20 ···················· ···················· ····················· ···················· ···················· ····················
W 3571 hCoV-19/England/ALDP-BE8F7F/2020|EPI_ISL_708888|20 ···················· ···················· ····················· ···················· ···················· ··A·················
W 3572 hCoV-19/England/MILK-BEE270/2020|EPI_ISL_708889|20 ···················· ·A·················· ····················· ···················· ···················· ··A·················
W 3573 hCoV-19/England/MILK-BEE52C/2020|EPI_ISL_708900|20 ···················· ···················· ····················· ···················· ···················· ····················
W 3574 hCoV-19/England/ALDP-BE959A/2020|EPI_ISL_708902|20 ···················· ···················· ····················· ···················· ···················· ····················
W 3575 hCoV-19/England/ALDP-BE9521/2020|EPI_ISL_708903|20 ···················· ···················· ····················· ···················· ···················· ····················
W 3576 hCoV-19/England/QEUH-BEB097/2020|EPI_ISL_708905|20 ···················· ···················· ····················· ···················· ···················· ····················
W 3577 hCoV-19/England/QEUH-BEAFC8/2020|EPI_ISL_708906|20 ···················· ···················· ····················· ···················· ···················· ····················
W 3578 hCoV-19/England/ALDP-BE914B/2020|EPI_ISL_708907|20 ···················· ···················· ····················· ···················· ···················· ····················
W 3579 hCoV-19/England/ALDP-BE8FE8/2020|EPI_ISL_708931|20 ···················· ···················· ····················· ···················· ···················· ····················
W 3580 hCoV-19/England/ALDP-BE903F/2020|EPI_ISL_708932|20 ···················· ···················· ····················· ···················· ···················· ····················
W 3581 hCoV-19/England/QEUH-BEB48C/2020|EPI_ISL_708933|20 ···················· ···················· ····················· ···················· ···················· ····················
W 3582 hCoV-19/England/ALDP-BE9372/2020|EPI_ISL_708935|20 ···················· ···················· ····················· ···················· ···················· ····················
W 3583 hCoV-19/England/ALDP-BE9E35/2020|EPI_ISL_708936|20 ···················· ···················· ····················· ···················· ···················· ····················
W 3584 hCoV-19/England/ALDP-BE91A5/2020|EPI_ISL_708937|20 ···················· ···················· ····················· ···················· ···················· ····················
W 3585 hCoV-19/USA/AZ-TG378641/2020|EPI_ISL_694630|2020-0 ···················· ···················· ····················· ···················· ···················· ····················
W 3586 hCoV-19/USA/AZ-TG378900/2020|EPI_ISL_694632|2020-0 ···················· ···················· ····················· ···················· ···················· ····················
W 3587 hCoV-19/USA/AZ-TG379182/2020|EPI_ISL_694635|2020-0 ···················· ···················· ····················· ···················· ···················· ····················
W 3588 hCoV-19/USA/AZ-TG379342/2020|EPI_ISL_694636|2020-0 ···················· ···················· ····················· ···················· ···················· ····················
W 3589 hCoV-19/USA/AZ-TG379388/2020|EPI_ISL_694637|2020-0 ···················· ···················· ····················· ···················· ···················· ····················
W 3590 hCoV-19/USA/AZ-TG379464/2020|EPI_ISL_694638|2020-0 ···················· ·N·················· ····················· ···················· ···················· ····················
W 3591 hCoV-19/USA/AZ-TG379485/2020|EPI_ISL_694639|2020-0 ···················· ···················· ····················· ···················· ···················· ····················
W 3592 hCoV-19/USA/AZ-TG379817/2020|EPI_ISL_694640|2020-0 ···················· ···················· ····················· ···················· ···················· ····················
W 3593 hCoV-19/USA/AZ-TG379993/2020|EPI_ISL_694641|2020-0 ···················· ·A·················· ····················· ···················· ···················· ····················
W 3594 hCoV-19/USA/AZ-TG379994/2020|EPI_ISL_694642|2020-0 ···················· ···················· ····················· ···················· ···················· ····················
W 3595 hCoV-19/USA/AZ-TG380005/2020|EPI_ISL_694643|2020-0 ···················· ···················· ····················· ···················· ···················· ····················
W 3596 hCoV-19/USA/AZ-TG380029/2020|EPI_ISL_694644|2020-0 ···················· ···················· ····················· ···················· ···················· ····················
W 3597 hCoV-19/USA/AZ-TG380033/2020|EPI_ISL_694645|2020-0 ···················· ·A·················· ····················· ···················· ···················· ····················
W 3598 hCoV-19/USA/AZ-TG380688/2020|EPI_ISL_694646|2020-0 ···················· ··N················· ····················· ···················· ···················· ····················
W 3599 hCoV-19/USA/AZ-TG381052/2020|EPI_ISL_694648|2020-0 ···················· ···················· ····················· ···················· ···················· ····················
W 3600 hCoV-19/USA/AZ-TG381162/2020|EPI_ISL_694649|2020-0 ···················· ···················· ····················· ···················· ···················· ····················
W 3601 hCoV-19/USA/AZ-TG381235/2020|EPI_ISL_694651|2020-0 ···················· ···················· ····················· ···················· ···················· ····················
W 3602 hCoV-19/USA/AZ-TG381245/2020|EPI_ISL_694652|2020-0 ···················· ···················· ····················· ···················· ···················· ····················
W 3603 hCoV-19/USA/AZ-TG381246/2020|EPI_ISL_694653|2020-0 ···················· ···················· ····················· ···················· ···················· ····················
W 3604 hCoV-19/USA/AZ-TG381248/2020|EPI_ISL_694654|2020-0 ···················· ···················· ····················· ···················· ···················· ····················
W 3605 hCoV-19/USA/AZ-TG381249/2020|EPI_ISL_694655|2020-0 ···················· ···················· ····················· ···················· ···················· ····················
W 3606 hCoV-19/USA/AZ-TG381250/2020|EPI_ISL_694656|2020-0 ···················· ···················· ····················· ···················· ···················· ····················
W 3607 hCoV-19/USA/AZ-TG381366/2020|EPI_ISL_694657|2020-0 ···················· ···················· ····················· ···················· ···················· ····················
W 3608 hCoV-19/USA/AZ-TG381367/2020|EPI_ISL_694658|2020-0 ···················· ···················· ····················· ···················· ···················· ····················
W 3609 hCoV-19/USA/AZ-TG381368/2020|EPI_ISL_694659|2020-0 ···················· ···················· ····················· ···················· ···················· ····················
W 3610 hCoV-19/USA/AZ-TG381370/2020|EPI_ISL_694660|2020-0 ···················· ···················· ····················· ···················· ···················· ····················
W 3611 hCoV-19/USA/AZ-TG381448/2020|EPI_ISL_694662|2020-0 ···················· ···················· ····················· ···················· ···················· ····················
W 3612 hCoV-19/USA/AZ-TG381593/2020|EPI_ISL_694663|2020-0 ···················· ···················· ····················· ···················· ···················· ····················
W 3613 hCoV-19/USA/AZ-TG381594/2020|EPI_ISL_694664|2020-0 ···················· ···················· ····················· ···················· ···················· ····················
W 3614 hCoV-19/USA/AZ-TG381595/2020|EPI_ISL_694665|2020-0 ···················· ···················· ····················· ···················· ···················· ····················
W 3615 hCoV-19/USA/AZ-TG381596/2020|EPI_ISL_694666|2020-0 ···················· ···················· ····················· ···················· ···················· ····················
W 3616 hCoV-19/USA/AZ-TG381597/2020|EPI_ISL_694667|2020-0 ···················· ·N·················· ····················· ···················· ···················· ····················
W 3617 hCoV-19/USA/AZ-TG381598/2020|EPI_ISL_694668|2020-0 ···················· ·N·················· ····················· ···················· ···················· ····················
W 3618 hCoV-19/USA/AZ-TG381610/2020|EPI_ISL_694669|2020-0 ···················· ·A·················· ····················· ···················· ···················· ····················
W 3619 hCoV-19/USA/AZ-TG381612/2020|EPI_ISL_694670|2020-0 ···················· ···················· ····················· ···················· ···················· ····················
W 3620 hCoV-19/USA/AZ-TG381613/2020|EPI_ISL_694671|2020-0 ···················· ···················· ····················· ···················· ···················· ····················
W 3621 hCoV-19/USA/AZ-TG381614/2020|EPI_ISL_694672|2020-0 ···················· ···················· ····················· ···················· ···················· ····················
W 3622 hCoV-19/USA/AZ-TG381616/2020|EPI_ISL_694673|2020-0 ···················· ···················· ····················· ···················· ···················· ····················
W 3623 hCoV-19/USA/AZ-TG382133/2020|EPI_ISL_694674|2020-0 ···················· ·N·················· ····················· ···················· ···················· ····················
W 3624 hCoV-19/USA/AZ-TG382137/2020|EPI_ISL_694675|2020-0 ···················· ···················· ····················· ···················· ···················· ····················
W 3625 hCoV-19/USA/AZ-TG382138/2020|EPI_ISL_694676|2020-0 ···················· ···················· ····················· ···················· ···················· ····················
W 3626 hCoV-19/USA/AZ-TG382139/2020|EPI_ISL_694677|2020-0 ···················· ···················· ····················· ···················· ···················· ····················
W 3627 hCoV-19/USA/AZ-TG382194/2020|EPI_ISL_694678|2020-0 ···················· ···················· ····················· ···················· ···················· ····················
W 3628 hCoV-19/USA/AZ-TG382195/2020|EPI_ISL_694679|2020-0 ···················· ···················· ····················· ···················· ···················· ····················
W 3629 hCoV-19/USA/AZ-TG382196/2020|EPI_ISL_694680|2020-0 ···················· ···················· ····················· ···················· ···················· ····················
W 3630 hCoV-19/USA/AZ-TG382198/2020|EPI_ISL_694681|2020-0 ···················· ···················· ····················· ···················· ···················· ····················
W 3631 hCoV-19/USA/AZ-TG382200/2020|EPI_ISL_694682|2020-0 ···················· ···················· ····················· ···················· ···················· ····················
W 3632 hCoV-19/USA/AZ-TG382285/2020|EPI_ISL_694683|2020-0 ···················· ···················· ····················· ···················· ···················· ····················
W 3633 hCoV-19/USA/AZ-TG382290/2020|EPI_ISL_694684|2020-0 ···················· ···················· ····················· ···················· ···················· ····················
W 3634 hCoV-19/USA/AZ-TG383297/2020|EPI_ISL_694687|2020-0 ···················· ···················· ····················· ···················· ···················· ····················
W 3635 hCoV-19/USA/AZ-TG383538/2020|EPI_ISL_694688|2020-0 ···················· ·A·················· ····················· ···················· ···················· ····················
W 3636 hCoV-19/USA/AZ-TG383704/2020|EPI_ISL_694689|2020-0 ···················· ···················· ····················· ···················· ···················· ····················

| | | | | | | | |
|---|---|---|---|---|---|---|---|
| W | 3637 hCoV-19/USA/AZ-TG383705/2020\|EPI_ISL_694690\|2020-0 | ···················· | ···················· | ···················· | ···················· | ···················· | ···················· |
| W | 3638 hCoV-19/USA/AZ-TG383720/2020\|EPI_ISL_694691\|2020-0 | ···················· | ···················· | ···················· | ···················· | ···················· | ···················· |
| W | 3639 hCoV-19/USA/AZ-TG384418/2020\|EPI_ISL_694692\|2020-0 | ···················· | ···················· | ···················· | ···················· | ···················· | ···················· |
| W | 3640 hCoV-19/USA/AZ-TG384422/2020\|EPI_ISL_694693\|2020-0 | ···················· | ···················· | ···················· | ···················· | ···················· | ···················· |
| W | 3641 hCoV-19/USA/AZ-TG384976/2020\|EPI_ISL_694695\|2020-0 | ···················· | ···················· | ···················· | ···················· | ···················· | ···················· |
| W | 3642 hCoV-19/USA/AZ-TG384982/2020\|EPI_ISL_694696\|2020-0 | ···················· | ···················· | ···················· | ···················· | ···················· | ···················· |
| W | 3643 hCoV-19/USA/AZ-TG388028/2020\|EPI_ISL_694697\|2020-0 | ···················· | ···················· | ···················· | ···················· | ···················· | ···················· |
| W | 3644 hCoV-19/USA/AZ-TG389333/2020\|EPI_ISL_694698\|2020-0 | ···················· | ···················· | ···················· | ···················· | ···················· | ···················· |
| W | 3645 hCoV-19/USA/AZ-TG389339/2020\|EPI_ISL_694699\|2020-0 | ···················· | ···················· | ···················· | ···················· | ···················· | ···················· |
| W | 3646 hCoV-19/USA/AZ-TG654313/2020\|EPI_ISL_695968\|2020-0 | ···················· | ···················· | ···················· | ···················· | ···················· | ···················· |
| W | 3647 hCoV-19/USA/AZ-TG654314/2020\|EPI_ISL_695969\|2020-0 | ···················· | ···················· | ···················· | ···················· | ···················· | ···················· |
| W | 3648 hCoV-19/USA/AZ-TG654315/2020\|EPI_ISL_695970\|2020-0 | ···················· | ···················· | ···················· | ···················· | ···················· | ···················· |
| W | 3649 hCoV-19/USA/AZ-TG654319/2020\|EPI_ISL_695972\|2020-0 | ···················· | ···················· | ···················· | ···················· | ···················· | ···················· |
| W | 3650 hCoV-19/USA/AZ-TG654322/2020\|EPI_ISL_695975\|2020-0 | ···················· | ···················· | ···················· | ···················· | ···················· | ···················· |
| W | 3651 hCoV-19/USA/AZ-TG654326/2020\|EPI_ISL_695976\|2020-0 | ···················· | ···················· | ···················· | ···················· | ···················· | ···················· |
| W | 3652 hCoV-19/USA/AZ-TG654332/2020\|EPI_ISL_695981\|2020-0 | ···················· | ···················· | ···················· | ···················· | ···················· | ···················· |
| W | 3653 hCoV-19/United Arab Emirates/1086/2020\|EPI_ISL_698 | ···················· | ···················· | ···················· | ···················· | ···················· | ···················· |
| W | 3654 hCoV-19/United Arab Emirates/1087/2020\|EPI_ISL_698 | ···················· | ···················· | ···················· | ···················· | ···················· | ···················· |
| W | 3655 hCoV-19/United Arab Emirates/1088/2020\|EPI_ISL_698 | ···················· | ···················· | ···················· | ···················· | ···················· | ···················· |
| W | 3656 hCoV-19/United Arab Emirates/1089/2020\|EPI_ISL_698 | ···················· | ···················· | ···················· | ···················· | ···················· | ···················· |
| W | 3657 hCoV-19/United Arab Emirates/1090/2020\|EPI_ISL_698 | ···················· | ···················· | ···················· | ···················· | ···················· | ···················· |
| W | 3658 hCoV-19/United Arab Emirates/1091/2020\|EPI_ISL_698 | ···················· | ···················· | ···················· | ···················· | ···················· | ···················· |
| W | 3659 hCoV-19/United Arab Emirates/1093/2020\|EPI_ISL_698 | ···················· | ···················· | ···················· | ···················· | ···················· | ···················· |
| W | 3660 hCoV-19/United Arab Emirates/1094/2020\|EPI_ISL_698 | ···················· | ···················· | ···················· | ···················· | ···················· | ···················· |
| W | 3661 hCoV-19/United Arab Emirates/1095/2020\|EPI_ISL_698 | ···················· | ···················· | ···················· | ···················· | ···················· | ···················· |
| W | 3662 hCoV-19/United Arab Emirates/1096/2020\|EPI_ISL_698 | ···················· | ···················· | ···················· | ···················· | ···················· | ···················· |
| W | 3663 hCoV-19/England/QEUH-BC47BD/2020\|EPI_ISL_703262\|20 | ···················· | ···················· | ···················· | ···················· | ···················· | ···················· |
| W | 3664 hCoV-19/England/QEUH-BC3EE6/2020\|EPI_ISL_703263\|20 | ···················· | ···················· | ···················· | ···················· | ···················· | ···················· |
| W | 3665 hCoV-19/England/ALDP-BCA5BD/2020\|EPI_ISL_703265\|20 | ···················· | ·A·················· | ···················· | ···················· | ···················· | ···················· |
| W | 3666 hCoV-19/England/ALDP-BCA939/2020\|EPI_ISL_703266\|20 | ···················· | ···················· | ···················· | ···················· | ···················· | ···················· |
| W | 3667 hCoV-19/England/ALDP-BCA571/2020\|EPI_ISL_703268\|20 | ···················· | ···················· | ···················· | ···················· | ···················· | ···················· |
| W | 3668 hCoV-19/England/QEUH-BC4708/2020\|EPI_ISL_703270\|20 | ···················· | ···················· | ···················· | ···················· | ···················· | ···················· |
| W | 3669 hCoV-19/England/ALDP-BCA6C9/2020\|EPI_ISL_703271\|20 | ···················· | ···················· | ···················· | ···················· | ···················· | ···················· |
| W | 3670 hCoV-19/England/ALDP-BCA0AD/2020\|EPI_ISL_703274\|20 | ···················· | ···················· | ···················· | ···················· | ···················· | ···················· |
| W | 3671 hCoV-19/England/ALDP-BCA3C2/2020\|EPI_ISL_703277\|20 | ···················· | ···················· | ···················· | ···················· | ···················· | ···················· |
| W | 3672 hCoV-19/England/QEUH-BC3EB9/2020\|EPI_ISL_703280\|20 | ···················· | ···················· | ···················· | ···················· | ···················· | ···················· |
| W | 3673 hCoV-19/England/ALDP-BCAB60/2020\|EPI_ISL_703282\|20 | ···················· | ···················· | ···················· | ···················· | ···················· | ···················· |
| W | 3674 hCoV-19/England/QEUH-BC45D1/2020\|EPI_ISL_703285\|20 | ···················· | ···················· | ···················· | ···················· | ···················· | ···················· |
| W | 3675 hCoV-19/England/QEUH-BC4313/2020\|EPI_ISL_703286\|20 | ···················· | ···················· | ···················· | ···················· | ···················· | ···················· |
| W | 3676 hCoV-19/England/QEUH-BC3FF2/2020\|EPI_ISL_703290\|20 | ···················· | ···················· | ···················· | ···················· | ···················· | ··A················· |
| W | 3677 hCoV-19/England/QEUH-BC454A/2020\|EPI_ISL_703291\|20 | ···················· | ·A·················· | ···················· | ···················· | ···················· | ···················· |
| W | 3678 hCoV-19/England/ALDP-BCA9C0/2020\|EPI_ISL_703293\|20 | ···················· | ···················· | ···················· | ···················· | ···················· | ···················· |
| W | 3679 hCoV-19/England/ALDP-BCABAC/2020\|EPI_ISL_703294\|20 | ···················· | ···················· | ···················· | ···················· | ···················· | ···················· |
| W | 3680 hCoV-19/England/QEUH-BC4647/2020\|EPI_ISL_703296\|20 | ···················· | ···················· | ···················· | ···················· | ···················· | ···················· |
| W | 3681 hCoV-19/England/ALDP-BCABCA/2020\|EPI_ISL_703298\|20 | ···················· | ···················· | ···················· | ···················· | ···················· | ···················· |
| W | 3682 hCoV-19/England/QEUH-A6206C/2020\|EPI_ISL_704594\|20 | ···················· | ···················· | ···················· | ···················· | ···················· | ···················· |
| W | 3683 hCoV-19/Scotland/CVR5974/2020\|EPI_ISL_704595\|2020- | ···················· | ···················· | ···················· | ···················· | ···················· | ···················· |
| W | 3684 hCoV-19/Scotland/CVR6031/2020\|EPI_ISL_704596\|2020- | ···················· | ···················· | ···················· | ···················· | ···················· | ···················· |
| W | 3685 hCoV-19/Scotland/CVR6032/2020\|EPI_ISL_704597\|2020- | ···················· | ···················· | ···················· | ···················· | ···················· | ···················· |
| W | 3686 hCoV-19/England/LIVE-DC4672/2020\|EPI_ISL_705900\|20 | ···················· | ···················· | ···················· | ···················· | ···················· | ···················· |
| W | 3687 hCoV-19/Norway/5461/2020\|EPI_ISL_708100\|2020-10-28 | ···················· | ···················· | ···················· | ···················· | ···················· | ···················· |
| W | 3688 hCoV-19/Norway/5464/2020\|EPI_ISL_708101\|2020-11-02 | ···················· | ···················· | ···················· | ···················· | ···················· | ···················· |
| W | 3689 hCoV-19/Norway/5466/2020\|EPI_ISL_708102\|2020-10-31 | ···················· | ···················· | ···················· | ···················· | ···················· | ···················· |
| W | 3690 hCoV-19/Norway/5467/2020\|EPI_ISL_708103\|2020-10-31 | ···················· | ···················· | ···················· | ···················· | ···················· | ···················· |
| W | 3691 hCoV-19/Norway/5470/2020\|EPI_ISL_708104\|2020-11-03 | ···················· | ···················· | ···················· | ···················· | ···················· | ···················· |
| W | 3692 hCoV-19/Norway/5472/2020\|EPI_ISL_708105\|2020-11-03 | ···················· | ···················· | ···················· | ···················· | ···················· | ···················· |
| W | 3693 hCoV-19/Norway/5487/2020\|EPI_ISL_708106\|2020-11-05 | ···················· | ···················· | ···················· | ···················· | ···················· | ···················· |
| W | 3694 hCoV-19/Norway/5489/2020\|EPI_ISL_708107\|2020-11-04 | ···················· | ···················· | ···················· | ···················· | ···················· | ···················· |
| W | 3695 hCoV-19/Norway/5493/2020\|EPI_ISL_708108\|2020-11-04 | ···················· | ···················· | ···················· | ···················· | ···················· | ···················· |
| W | 3696 hCoV-19/Norway/5494/2020\|EPI_ISL_708109\|2020-11-06 | ···················· | ···················· | ···················· | ···················· | ···················· | ···················· |
| W | 3697 hCoV-19/Norway/5535/2020\|EPI_ISL_708110\|2020-11-02 | ···················· | ·A·················· | ···················· | ···················· | ···················· | ··A················· |
| W | 3698 hCoV-19/Norway/5536/2020\|EPI_ISL_708111\|2020-11-03 | ···················· | ···················· | ···················· | ···················· | ···················· | ···················· |
| W | 3699 hCoV-19/Norway/5537/2020\|EPI_ISL_708112\|2020-11-03 | ···················· | ···················· | ···················· | ···················· | ···················· | ···················· |
| W | 3700 hCoV-19/Norway/5538/2020\|EPI_ISL_708113\|2020-11-04 | ···················· | ···················· | ···················· | ···················· | ···················· | ···················· |
| W | 3701 hCoV-19/Norway/5539/2020\|EPI_ISL_708114\|2020-11-04 | ···················· | ·A·················· | ···················· | ···················· | ···················· | ··A················· |
| W | 3702 hCoV-19/Norway/5543/2020\|EPI_ISL_708115\|2020-11-12 | ···················· | ···················· | ···················· | ···················· | ···················· | ···················· |
| W | 3703 hCoV-19/Norway/5544/2020\|EPI_ISL_708116\|2020-11-06 | ···················· | ·A·················· | ···················· | ···················· | ···················· | ··A················· |
| W | 3704 hCoV-19/Norway/5545/2020\|EPI_ISL_708117\|2020-11-06 | ···················· | ···················· | ···················· | ···················· | ···················· | ···················· |
| W | 3705 hCoV-19/Norway/5549/2020\|EPI_ISL_708118\|2020-11-07 | ···················· | ···················· | ···················· | ···················· | ···················· | ···················· |
| W | 3706 hCoV-19/Norway/5550/2020\|EPI_ISL_708119\|2020-11-10 | ···················· | ···················· | ···················· | ···················· | ···················· | ···················· |
| W | 3707 hCoV-19/Norway/5557/2020\|EPI_ISL_708120\|2020-11-07 | ···················· | ·A·················· | ···················· | ···················· | ···················· | ··A················· |
| W | 3708 hCoV-19/Norway/5560/2020\|EPI_ISL_708121\|2020-11-08 | ···················· | ·A·················· | ···················· | ···················· | ···················· | ··A················· |
| W | 3709 hCoV-19/Norway/5606/2020\|EPI_ISL_708122\|2020-11-09 | ···················· | ·A·················· | ···················· | ···················· | ···················· | ··A················· |
| W | 3710 hCoV-19/Norway/5607/2020\|EPI_ISL_708123\|2020-11-09 | ···················· | ·A·················· | ···················· | ···················· | ···················· | ··A················· |
| W | 3711 hCoV-19/Norway/5608/2020\|EPI_ISL_708124\|2020-11-09 | ···················· | ·A·················· | ···················· | ···················· | ···················· | ··A················· |
| W | 3712 hCoV-19/Norway/5609/2020\|EPI_ISL_708125\|2020-11-09 | ···················· | ·A·················· | ···················· | ···················· | ···················· | ··A················· |
| W | 3716 hCoV-19/Norway/5620/2020\|EPI_ISL_708129\|2020-11-10 | ···················· | ·A·················· | ···················· | ···················· | ···················· | ··A················· |
| W | 3718 hCoV-19/Norway/5641/2020\|EPI_ISL_708131\|2020-11-14 | ···················· | ···················· | ···················· | ···················· | ······T············· | ···················· |
| W | 3719 hCoV-19/Norway/5642/2020\|EPI_ISL_708132\|2020-11-09 | ···················· | ···················· | ···················· | ···················· | ···················· | ···················· |
| W | 3720 hCoV-19/Norway/5643/2020\|EPI_ISL_708133\|2020-11-10 | ···················· | ···················· | ···················· | ···················· | ···················· | ···················· |

| | | | | | | | | |
|---|---|---|---|---|---|---|---|---|
| W | 3721 | hCoV-19/Norway/5644/2020\|EPI_ISL_708134\|2020-11-10 | ···················· | ···················· | ····················· | ···················· | ···················· | ···················· |
| W | 3722 | hCoV-19/Norway/5645/2020\|EPI_ISL_708135\|2020-11-11 | ···················· | ···················· | ····················· | ···················· | ······T············· | ···················· |
| W | 3723 | hCoV-19/Norway/5654/2020\|EPI_ISL_708136\|2020-11-17 | ···················· | ···················· | ····················· | ···················· | ···················· | ···················· |
| W | 3724 | hCoV-19/Norway/5656/2020\|EPI_ISL_708137\|2020-11-17 | ···················· | ·A·················· | ····················· | ···················· | ···················· | ···A················ |
| W | 3725 | hCoV-19/Norway/5657/2020\|EPI_ISL_708138\|2020-11-17 | ···················· | ···················· | ····················· | ···················· | ···················· | ···················· |
| W | 3726 | hCoV-19/Norway/5658/2020\|EPI_ISL_708139\|2020-11-17 | ···················· | ·A·················· | ····················· | ···················· | ···················· | ···A················ |
| W | 3727 | hCoV-19/Norway/5660/2020\|EPI_ISL_708140\|2020-11-17 | ···················· | ·A·················· | ····················· | ···················· | ···················· | ···A················ |
| W | 3728 | hCoV-19/Norway/5663/2020\|EPI_ISL_708141\|2020-10-30 | ···················· | ···················· | ····················· | ···················· | ······T············· | ···················· |
| W | 3729 | hCoV-19/Norway/5705/2020\|EPI_ISL_708143\|2020-11-09 | ···················· | ···················· | ····················· | ···················· | ···················· | ···················· |
| W | 3730 | hCoV-19/Norway/5706/2020\|EPI_ISL_708144\|2020-11-09 | ···················· | ···················· | ····················· | ···················· | ···················· | ···················· |
| W | 3731 | hCoV-19/Norway/5707/2020\|EPI_ISL_708145\|2020-11-08 | ···················· | ·A·················· | ····················· | ···················· | ···················· | ···A················ |
| W | 3732 | hCoV-19/Norway/5708/2020\|EPI_ISL_708146\|2020-11-09 | ···················· | ···················· | ····················· | ···················· | ···················· | ···················· |
| W | 3733 | hCoV-19/Norway/5710/2020\|EPI_ISL_708147\|2020-11-10 | ···················· | ·A·················· | ····················· | ···················· | ···················· | ···A················ |
| W | 3734 | hCoV-19/Norway/5711/2020\|EPI_ISL_708148\|2020-11-09 | ···················· | ···················· | ····················· | ···················· | ···················· | ···················· |
| W | 3735 | hCoV-19/Norway/5712/2020\|EPI_ISL_708149\|2020-11-10 | ···················· | ·A·················· | ····················· | ···················· | ···················· | ···A················ |
| W | 3736 | hCoV-19/Norway/5782/2020\|EPI_ISL_708170\|2020-11-19 | ···················· | ···················· | ····················· | ···················· | ···················· | ···················· |
| W | 3737 | hCoV-19/Norway/5789/2020\|EPI_ISL_708171\|2020-11-14 | ···················· | ···················· | ····················· | ···················· | ···················· | ···················· |
| W | 3738 | hCoV-19/Norway/5790/2020\|EPI_ISL_708172\|2020-11-17 | ···················· | ·A·················· | ····················· | ···················· | ···················· | ···A················ |
| W | 3739 | hCoV-19/Norway/5791/2020\|EPI_ISL_708173\|2020-11-17 | ···················· | ·A·················· | ····················· | ···················· | ···················· | ···A················ |
| W | 3740 | hCoV-19/Norway/5792/2020\|EPI_ISL_708174\|2020-11-18 | ···················· | ···················· | ····················· | ···················· | ···················· | ···················· |
| W | 3741 | hCoV-19/Norway/5793/2020\|EPI_ISL_708175\|2020-11-18 | ···················· | ···················· | ····················· | ···················· | ······T············· | ···················· |
| W | 3742 | hCoV-19/Norway/5794/2020\|EPI_ISL_708176\|2020-11-09 | ···················· | ·A·················· | ····················· | ···················· | ···················· | ···A················ |
| W | 3743 | hCoV-19/Norway/5796/2020\|EPI_ISL_708177\|2020-11-09 | ···················· | ···················· | ····················· | ···················· | ···················· | ···················· |
| W | 3744 | hCoV-19/Norway/5797/2020\|EPI_ISL_708178\|2020-11-10 | ···················· | ···················· | ····················· | ···················· | ···················· | ···················· |
| W | 3745 | hCoV-19/Norway/5798/2020\|EPI_ISL_708179\|2020-11-14 | ···················· | ···················· | ····················· | ···················· | ···················· | ···················· |
| W | 3746 | hCoV-19/England/QEUH-BEB088/2020\|EPI_ISL_708910\|20 | ···················· | ···················· | ····················· | ···················· | ···················· | ···················· |
| W | 3747 | hCoV-19/England/ALDP-BE93BE/2020\|EPI_ISL_708911\|20 | ···················· | ···················· | ····················· | ···················· | ···················· | ···················· |
| W | 3748 | hCoV-19/England/MILK-BEE42F/2020\|EPI_ISL_708912\|20 | ···················· | ···················· | ····················· | ···················· | ······T············· | ···················· |
| W | 3749 | hCoV-19/England/MILK-BEE51D/2020\|EPI_ISL_708913\|20 | ···················· | ·A·················· | ····················· | ···················· | ···················· | ···················· |
| W | 3750 | hCoV-19/England/ALDP-BE93AF/2020\|EPI_ISL_708914\|20 | ···················· | ···················· | ····················· | ···················· | ···················· | ···················· |
| W | 3751 | hCoV-19/England/MILK-BEE28F/2020\|EPI_ISL_708916\|20 | ···················· | ···················· | ····················· | ···················· | ···················· | ···················· |
| W | 3752 | hCoV-19/England/ALDP-BE8EAF/2020\|EPI_ISL_708917\|20 | ···················· | ···················· | ····················· | ···················· | ···················· | ···················· |
| W | 3753 | hCoV-19/England/ALDP-BE96B5/2020\|EPI_ISL_708919\|20 | ···················· | ···················· | ····················· | ···················· | ···················· | ···················· |
| W | 3754 | hCoV-19/England/MILK-BEE47A/2020\|EPI_ISL_708920\|20 | ···················· | ···················· | ····················· | ···················· | ···················· | ···················· |
| W | 3755 | hCoV-19/England/QEUH-BEB440/2020\|EPI_ISL_708922\|20 | ···················· | ···················· | ····················· | ···················· | ···················· | ···················· |
| W | 3756 | hCoV-19/England/ALDP-BE91C3/2020\|EPI_ISL_708923\|20 | ···················· | ···················· | ····················· | ···················· | ···················· | ···················· |
| W | 3757 | hCoV-19/England/ALDP-BE8F42/2020\|EPI_ISL_708924\|20 | ···················· | ···················· | ····················· | ···················· | ···················· | ···A················ |
| W | 3758 | hCoV-19/England/ALDP-BE9512/2020\|EPI_ISL_708929\|20 | ···················· | ···················· | ····················· | ···················· | ···················· | ···················· |
| W | 3759 | hCoV-19/England/ALDP-BE980A/2020\|EPI_ISL_708940\|20 | ···················· | ···················· | ····················· | ···················· | ···················· | ···················· |
| W | 3760 | hCoV-19/England/ALDP-BE8E27/2020\|EPI_ISL_708942\|20 | ···················· | ···················· | ····················· | ···················· | ······T············· | ···················· |
| W | 3761 | hCoV-19/England/ALDP-BE8EFA/2020\|EPI_ISL_708943\|20 | ···················· | ···················· | ····················· | ···················· | ···················· | ···A················ |
| W | 3762 | hCoV-19/England/ALDP-BE9363/2020\|EPI_ISL_708946\|20 | ···················· | ···················· | ····················· | ···················· | ···················· | ···················· |
| W | 3763 | hCoV-19/England/ALDP-BE9248/2020\|EPI_ISL_708947\|20 | ···················· | ···················· | ····················· | ···················· | ···················· | ···················· |
| W | 3764 | hCoV-19/England/ALDP-BE9020/2020\|EPI_ISL_708949\|20 | ···················· | ···················· | ····················· | ···················· | ······T············· | ···················· |
| W | 3765 | hCoV-19/England/ALDP-C06147/2020\|EPI_ISL_709401\|20 | ···················· | ···················· | ····················· | ···················· | ···················· | ···················· |
| W | 3766 | hCoV-19/England/QEUH-C14674/2020\|EPI_ISL_709403\|20 | ···················· | ···················· | ····················· | ···················· | ···················· | ···················· |
| W | 3767 | hCoV-19/England/QEUH-C14665/2020\|EPI_ISL_709404\|20 | ···················· | ···················· | ····················· | ···················· | ······T············· | ···················· |
| W | 3768 | hCoV-19/England/QEUH-C1461A/2020\|EPI_ISL_709406\|20 | ···················· | ···················· | ····················· | ···················· | ······T············· | ···················· |
| W | 3769 | hCoV-19/England/QEUH-C16038/2020\|EPI_ISL_709407\|20 | ···················· | ·A·················· | ····················· | ···················· | ···················· | ···················· |
| W | 3770 | hCoV-19/England/QEUH-C15998/2020\|EPI_ISL_709409\|20 | ···················· | ···················· | ····················· | ···················· | ···················· | ···················· |
| W | 3771 | hCoV-19/England/ALDP-C06684/2020\|EPI_ISL_709410\|20 | ···················· | ···················· | ····················· | ···················· | ···················· | ···················· |
| W | 3772 | hCoV-19/England/ALDP-C063E7/2020\|EPI_ISL_709414\|20 | ···················· | ···················· | ····················· | ···················· | ···················· | ···················· |
| W | 3773 | hCoV-19/England/ALDP-C06745/2020\|EPI_ISL_709415\|20 | ···················· | ···················· | ····················· | ···················· | ···················· | ···················· |
| W | 3774 | hCoV-19/England/ALDP-C060D1/2020\|EPI_ISL_709416\|20 | ···················· | ···················· | ····················· | ···················· | ···················· | ···················· |
| W | 3775 | hCoV-19/England/QEUH-C14E6D/2020\|EPI_ISL_709417\|20 | ···················· | ···················· | ····················· | ···················· | ···················· | ···················· |
| W | 3776 | hCoV-19/England/ALDP-C063C9/2020\|EPI_ISL_709418\|20 | ···················· | ···················· | ····················· | ···················· | ···················· | ···················· |
| W | 3777 | hCoV-19/England/QEUH-C16056/2020\|EPI_ISL_709419\|20 | ···················· | ···················· | ····················· | ···················· | ···················· | ···················· |
| W | 3778 | hCoV-19/England/ALDP-C065E1/2020\|EPI_ISL_709421\|20 | ···················· | ···················· | ····················· | ···················· | ···················· | ···················· |
| W | 3779 | hCoV-19/England/QEUH-C1496C/2020\|EPI_ISL_709423\|20 | ···················· | ···················· | ····················· | ···················· | ···················· | ···················· |
| W | 3780 | hCoV-19/England/QEUH-C14E03/2020\|EPI_ISL_709424\|20 | ···················· | ·A·················· | ····················· | ···················· | ···················· | ···A················ |
| W | 3781 | hCoV-19/England/ALDP-C06411/2020\|EPI_ISL_709425\|20 | ···················· | ···················· | ····················· | ···················· | ···················· | ···················· |
| W | 3782 | hCoV-19/England/QEUH-C1643C/2020\|EPI_ISL_709426\|20 | ···················· | ···················· | ····················· | ···················· | ···················· | ···A················ |
| W | 3783 | hCoV-19/England/ALDP-C062CC/2020\|EPI_ISL_709427\|20 | ···················· | ···················· | ····················· | ···················· | ···················· | ···················· |
| W | 3784 | hCoV-19/England/ALDP-C066A2/2020\|EPI_ISL_709430\|20 | ···················· | ···················· | ····················· | ···················· | ···················· | ···················· |
| W | 3785 | hCoV-19/England/ALDP-C05F8A/2020\|EPI_ISL_709433\|20 | ···················· | ···················· | ····················· | ···················· | ···················· | ···················· |
| W | 3786 | hCoV-19/England/QEUH-C1493F/2020\|EPI_ISL_709436\|20 | ···················· | ·A·················· | ····················· | ···················· | ···················· | ···················· |
| W | 3787 | hCoV-19/England/ALDP-C06086/2020\|EPI_ISL_709437\|20 | ···················· | ···················· | ····················· | ···················· | ···················· | ···················· |
| W | 3788 | hCoV-19/England/QEUH-C1645A/2020\|EPI_ISL_709438\|20 | ···················· | ···················· | ····················· | ···················· | ···················· | ···················· |
| W | 3789 | hCoV-19/England/QEUH-C14A0F/2020\|EPI_ISL_709439\|20 | ···················· | ···················· | ····················· | ···················· | ···················· | ···················· |
| W | 3790 | hCoV-19/England/QEUH-C14A3C/2020\|EPI_ISL_709440\|20 | ···················· | ·A·················· | ····················· | ···················· | ···················· | ···A················ |
| W | 3791 | hCoV-19/England/QEUH-C14BA2/2020\|EPI_ISL_709441\|20 | ···················· | ···················· | ····················· | ···················· | ···················· | ···················· |
| W | 3792 | hCoV-19/England/QEUH-C149E4/2020\|EPI_ISL_709443\|20 | ···················· | ···················· | ····················· | ···················· | ···················· | ···················· |
| W | 3793 | hCoV-19/England/ALDP-C067CD/2020\|EPI_ISL_709444\|20 | ···················· | ···················· | ····················· | ···················· | ···················· | ···················· |
| W | 3794 | hCoV-19/England/ALDP-C06D70/2020\|EPI_ISL_709445\|20 | ···················· | ···················· | ····················· | ···················· | ···················· | ···················· |
| W | 3795 | hCoV-19/England/QEUH-C15FB4/2020\|EPI_ISL_709446\|20 | ···················· | ···················· | ····················· | ···················· | ···················· | ···················· |
| W | 3796 | hCoV-19/England/ALDP-C05FC6/2020\|EPI_ISL_709447\|20 | ···················· | ···················· | ····················· | ···················· | ···················· | ···················· |
| W | 3797 | hCoV-19/England/ALDP-C069D6/2020\|EPI_ISL_709448\|20 | ···················· | ···················· | ····················· | ···················· | ···················· | ···················· |
| W | 3798 | hCoV-19/England/QEUH-C1497B/2020\|EPI_ISL_709450\|20 | ···················· | ···················· | ····················· | ···················· | ···················· | ···················· |
| W | 3799 | hCoV-19/England/ALDP-C06192/2020\|EPI_ISL_709451\|20 | ···················· | ···················· | ····················· | ···················· | ···················· | ···················· |
| W | 3800 | hCoV-19/England/ALDP-C0601D/2020\|EPI_ISL_709452\|20 | ···················· | ···················· | ····················· | ···················· | ···················· | ···················· |

W 3801 hCoV-19/England/QEUH-C15989/2020|EPI_ISL_709453|20 ···················· ···················· ···················· ···················· ···················· ····················

W 3802 hCoV-19/England/QEUH-C14E7C/2020|EPI_ISL_709454|20 ···················· ···················· ···················· ···················· ···················· ····················

W 3803 hCoV-19/England/QEUH-C15FF0/2020|EPI_ISL_709456|20 ···················· ···················· ···················· ···················· ···················· ····················

W 3804 hCoV-19/England/QEUH-C14BFD/2020|EPI_ISL_709457|20 ···················· ···················· ···················· ···················· ···················· ····················

W 3805 hCoV-19/England/ALDP-C060FF/2020|EPI_ISL_709460|20 ···················· ···················· ···················· ···················· ···················· ····················

W 3806 hCoV-19/England/ALDP-C0645D/2020|EPI_ISL_709461|20 ···················· ···················· ···················· ···················· ···················· ····················

W 3807 hCoV-19/England/QEUH-C16496/2020|EPI_ISL_709464|20 ···················· ···················· ···················· ···················· ···················· ····················

W 3808 hCoV-19/England/QEUH-C14BEE/2020|EPI_ISL_709465|20 ···················· ···················· ···················· ···················· ······T············· ····················

W 3809 hCoV-19/England/QEUH-C146CF/2020|EPI_ISL_709466|20 ···················· ···················· ···················· ···················· ···················· ····················

W 3810 hCoV-19/England/ALDP-C0602C/2020|EPI_ISL_709468|20 ···················· ···················· ···················· ···················· ···················· ····················

W 3811 hCoV-19/England/QEUH-C17F0D/2020|EPI_ISL_709469|20 ···················· ···················· ···················· ···················· ···················· ····················

W 3812 hCoV-19/England/ALDP-C06903/2020|EPI_ISL_709470|20 ···················· ···················· ···················· ···················· ···················· ····················

W 3813 hCoV-19/England/ALDP-C0655A/2020|EPI_ISL_709471|20 ···················· ···················· ···················· ···················· ···················· ····················

W 3814 hCoV-19/England/QEUH-C1494E/2020|EPI_ISL_709472|20 ···················· ···················· ···················· ···················· ···················· ····················

W 3815 hCoV-19/England/QEUH-C163F3/2020|EPI_ISL_709474|20 ···················· ···················· ···················· ···················· ···················· ····················

W 3816 hCoV-19/England/ALDP-C06235/2020|EPI_ISL_709475|20 ···················· ···················· ···················· ···················· ···················· ····················

W 3817 hCoV-19/England/ALDP-C06587/2020|EPI_ISL_709477|20 ···················· ···················· ···················· ···················· ···················· ····················

W 3818 hCoV-19/England/ALDP-C062F9/2020|EPI_ISL_709478|20 ···················· ···················· ···················· ···················· ···················· ····················

W 3819 hCoV-19/England/ALDP-C06244/2020|EPI_ISL_709479|20 ···················· ···················· ···················· ···················· ···················· ····················

W 3820 hCoV-19/USA/TX-DSHS-1748/2020|EPI_ISL_710400|2020- ···················· ····T··············· ···················· ···················· ···················· ····················

W 3821 hCoV-19/USA/TX-DSHS-1753/2020|EPI_ISL_710401|2020- ···················· ···················· ···················· ···················· ···················· ····················

W 3822 hCoV-19/USA/TX-DSHS-1754/2020|EPI_ISL_710402|2020- ···················· ···················· ···················· ···················· ···················· ····················

W 3823 hCoV-19/USA/TX-DSHS-1755/2020|EPI_ISL_710403|2020- ···················· ···················· ···················· ···················· ···················· ····················

W 3824 hCoV-19/USA/TX-DSHS-1757/2020|EPI_ISL_710404|2020- ···················· ···················· ···················· ···················· ···················· ····················

W 3825 hCoV-19/USA/TX-DSHS-1759/2020|EPI_ISL_710405|2020- ···················· ·A·················· ···················· ···················· ···················· ····················

W 3826 hCoV-19/USA/TX-DSHS-1762/2020|EPI_ISL_710406|2020- ···················· ···················· ···················· ···················· ···················· ····················

W 3827 hCoV-19/USA/TX-DSHS-1764/2020|EPI_ISL_710407|2020- ···················· ·A·················· ···················· ···················· ···················· ····················

W 3828 hCoV-19/USA/TX-DSHS-1766/2020|EPI_ISL_710408|2020- ···················· ···················· ···················· ···················· ···················· ····················

W 3829 hCoV-19/USA/TX-DSHS-1767/2020|EPI_ISL_710409|2020- ···················· ·············A······ ···················· ···················· ···················· ····················

W 3830 hCoV-19/USA/TX-DSHS-1770/2020|EPI_ISL_710410|2020- ···················· ···················· ···················· ···················· ···················· ····················

W 3831 hCoV-19/USA/TX-DSHS-1771/2020|EPI_ISL_710411|2020- ···················· ···················· ···················· ···················· ···················· ····················

W 3832 hCoV-19/USA/TX-DSHS-1772/2020|EPI_ISL_710412|2020- ···················· ······A············· ···················· ···················· ···················· ····················

W 3833 hCoV-19/USA/TX-DSHS-1773/2020|EPI_ISL_710413|2020- ···················· ···················· ···················· ···················· ···················· ····················

W 3834 hCoV-19/USA/TX-DSHS-1774/2020|EPI_ISL_710414|2020- ···················· ···················· ···················· ···················· ···················· ····················

W 3835 hCoV-19/USA/TX-DSHS-1728/2020|EPI_ISL_710415|2020- ···················· ···················· ···················· ···················· ···················· ····················

W 3836 hCoV-19/USA/CA-LACPHL-AA00025/2020|EPI_ISL_710430| ···················· ···················· ···················· ···················· ···················· ····················

W 3837 hCoV-19/USA/CA-LACPHL-AA00029/2020|EPI_ISL_710431| ···················· ···················· ···················· ···················· ···················· ····················

W 3838 hCoV-19/USA/CA-LACPHL-AA00032/2020|EPI_ISL_710432| ···················· ···················· ···················· ···················· ···················· ····················

W 3839 hCoV-19/Spain/AS-HUCA-232025446/2020|EPI_ISL_71049 ···················· ···················· ···················· ···················· ···················· ····················

W 3840 hCoV-19/Spain/AS-HUCA-232039084/2020|EPI_ISL_71049 ···················· ···················· ···················· ···················· ···················· ····················

W 3841 hCoV-19/USA/AZ-TG375454/2020|EPI_ISL_694603|2020-0 ···················· ···················· ···················· ···················· ···················· ····················

W 3842 hCoV-19/USA/AZ-TG375628/2020|EPI_ISL_694604|2020-0 ···················· ···················· ···················· ···················· ···················· ····················

W 3843 hCoV-19/USA/AZ-TG375921/2020|EPI_ISL_694607|2020-0 ···················· ···················· ···················· ···················· ···················· ····················

W 3844 hCoV-19/USA/AZ-TG375955/2020|EPI_ISL_694608|2020-0 ···················· ·A·················· ···················· ···················· ···················· ····················

W 3845 hCoV-19/USA/AZ-TG376112/2020|EPI_ISL_694612|2020-0 ···················· ·A·················· ···················· ···················· ···················· ····················

W 3846 hCoV-19/USA/AZ-TG376426/2020|EPI_ISL_694615|2020-0 ···················· ·A·················· ···················· ···················· ···················· ····················

W 3847 hCoV-19/USA/AZ-TG377242/2020|EPI_ISL_694616|2020-0 ···················· ···················· ···················· ···················· ···················· ····················

W 3848 hCoV-19/USA/AZ-TG377243/2020|EPI_ISL_694617|2020-0 ···················· ···················· ···················· ···················· ···················· ····················

W 3849 hCoV-19/USA/AZ-TG377253/2020|EPI_ISL_694619|2020-0 ···················· ···················· ···················· ···················· ···················· ····················

W 3850 hCoV-19/USA/AZ-TG377346/2020|EPI_ISL_694621|2020-0 ···················· ·A·················· ···················· ···················· ···················· ····················

W 3851 hCoV-19/USA/AZ-TG377347/2020|EPI_ISL_694622|2020-0 ···················· ·A·················· ···················· ···················· ···················· ····················

W 3852 hCoV-19/USA/AZ-TG377907/2020|EPI_ISL_694625|2020-0 ···················· ·A·················· ···················· ···················· ···················· ····················

W 3853 hCoV-19/USA/AZ-TG377909/2020|EPI_ISL_694626|2020-0 ···················· ···················· ···················· ···················· ···················· ····················

W 3854 hCoV-19/USA/AZ-TG378380/2020|EPI_ISL_694627|2020-0 ···················· ·A·················· ···················· ···················· ···················· ····················

W 3855 hCoV-19/USA/AZ-TG648723/2020|EPI_ISL_695908|2020-0 ···················· ···················· ···················· ···················· ···················· ····················

W 3856 hCoV-19/USA/AZ-TG648724/2020|EPI_ISL_695909|2020-0 ···················· ···················· ···················· ···················· ···················· ····················

W 3857 hCoV-19/USA/AZ-TG648726/2020|EPI_ISL_695911|2020-0 ···················· ···················· ···················· ···················· ···················· ····················

W 3858 hCoV-19/USA/AZ-TG648727/2020|EPI_ISL_695912|2020-0 ···················· ···················· ···················· ···················· ···················· ····················

W 3859 hCoV-19/USA/AZ-TG648728/2020|EPI_ISL_695913|2020-0 ···················· ···················· ···················· ···················· ···················· ····················

W 3860 hCoV-19/USA/AZ-TG648732/2020|EPI_ISL_695916|2020-0 ···················· ···················· ···················· ···················· ···················· ····················

W 3861 hCoV-19/USA/AZ-TG648734/2020|EPI_ISL_695917|2020-0 ···················· ···················· ···················· ···················· ·NN················· ····················

W 3862 hCoV-19/USA/AZ-TG648737/2020|EPI_ISL_695920|2020-0 ···················· ···················· ···················· ···················· ···················· ····················

W 3863 hCoV-19/USA/AZ-TG648738/2020|EPI_ISL_695921|2020-0 ···················· ···················· ···················· ···················· ···················· ····················

W 3864 hCoV-19/USA/AZ-TG648741/2020|EPI_ISL_695922|2020-0 ···················· ···················· ···················· ···················· ···················· ····················

W 3865 hCoV-19/USA/AZ-TG648749/2020|EPI_ISL_695926|2020-0 ···················· ···················· ···················· ···················· ···················· ····················

W 3866 hCoV-19/USA/AZ-TG648750/2020|EPI_ISL_695927|2020-0 ···················· ···················· ···················· ···················· ···················· ····················

W 3867 hCoV-19/USA/AZ-TG648768/2020|EPI_ISL_695939|2020-0 ···················· ···················· ···················· ···················· ···················· ····················

W 3868 hCoV-19/USA/AZ-TG648774/2020|EPI_ISL_695944|2020-0 ···················· ···················· ···················· ···················· ···················· ····················

W 3869 hCoV-19/USA/AZ-TG648777/2020|EPI_ISL_695945|2020-0 ···················· ···················· ···················· ···················· ···················· ····················

W 3870 hCoV-19/USA/AZ-TG654291/2020|EPI_ISL_695948|2020-0 ···················· ···················· ···················· ···················· ···················· ····················

W 3871 hCoV-19/USA/AZ-TG654301/2020|EPI_ISL_695957|2020-0 ···················· ···················· ···················· ···················· ···················· ····················

W 3872 hCoV-19/USA/AZ-TG654304/2020|EPI_ISL_695959|2020-0 ···················· ···················· ···················· ···················· ···················· ····················

W 3873 hCoV-19/Canada/ON-UHTC_0390/2020|EPI_ISL_698100|20 ···················· ···················· ···················· ···················· ···················· ····················

W 3874 hCoV-19/Canada/ON-UHTC_0383/2020|EPI_ISL_698101|20 ···················· ·A·················· ···················· ···················· ···················· ···A················

W 3875 hCoV-19/Canada/ON-UHTC_0386/2020|EPI_ISL_698102|20 ···················· ·A·················· ···················· ···················· ···················· ···A················

W 3876 hCoV-19/Canada/ON-UHTC_0381/2020|EPI_ISL_698103|20 ···················· ···················· ···················· ···················· ···················· ····················

W 3877 hCoV-19/Canada/ON-UHTC_0402/2020|EPI_ISL_698104|20 ···················· ···················· ···················· ···················· ···················· ····················

W 3878 hCoV-19/United Arab Emirates/1030/2020|EPI_ISL_698 ···················· ···················· ···················· ···················· ···················· ····················

W 3879 hCoV-19/United Arab Emirates/1031/2020|EPI_ISL_698 ···················· ···················· ···················· ···················· ···················· ····················

W 3880 hCoV-19/United Arab Emirates/1032/2020|EPI_ISL_698 ···················· ···················· ···················· ···················· ···················· ····················

W 3881 hCoV-19/United Arab Emirates/1033/2020|EPI_ISL_698 ···················· ···················· ···················· ···················· ···················· ····················
W 3882 hCoV-19/United Arab Emirates/1034/2020|EPI_ISL_698 ···················· ···················· ···················· ···················· ···················· ····················
W 3883 hCoV-19/United Arab Emirates/1035/2020|EPI_ISL_698 ···················· ···················· ···················· ···················· ···················· ····················
W 3884 hCoV-19/United Arab Emirates/1038/2020|EPI_ISL_698 ···················· ···················· ···················· ···················· ···················· ····················
W 3885 hCoV-19/United Arab Emirates/1039/2020|EPI_ISL_698 ···················· ···················· ···················· ···················· ···················· ····················
W 3886 hCoV-19/United Arab Emirates/1040/2020|EPI_ISL_698 ···················· ···················· ···················· ···················· ···················· ····················
W 3887 hCoV-19/United Arab Emirates/1041/2020|EPI_ISL_698 ···················· ···················· ···················· ···················· ···················· ····················
W 3888 hCoV-19/United Arab Emirates/0134/2020|EPI_ISL_698 ···················· ···················· ···················· ···················· ···················· ····················
W 3889 hCoV-19/United Arab Emirates/0135/2020|EPI_ISL_698 ···················· ···················· ···················· ···················· ···················· ····················
W 3890 hCoV-19/United Arab Emirates/0222/2020|EPI_ISL_698 ···················· ···················· ···················· ···················· ···················· ····················
W 3891 hCoV-19/United Arab Emirates/0225/2020|EPI_ISL_698 ···················· ···················· ···················· ···················· ···················· ····················
W 3892 hCoV-19/United Arab Emirates/0235/2020|EPI_ISL_698 ···················· ···················· ···················· ···················· ···················· ····················
W 3893 hCoV-19/United Arab Emirates/0309/2020|EPI_ISL_698 ···················· ···················· ···················· ···················· ···················· ····················
W 3894 hCoV-19/United Arab Emirates/0421/2020|EPI_ISL_698 ···················· ···················· ···················· ···················· ···················· ····················
W 3895 hCoV-19/United Arab Emirates/0565/2020|EPI_ISL_698 ···················· ···················· ···················· ···················· ···················· ····················
W 3896 hCoV-19/United Arab Emirates/0604/2020|EPI_ISL_698 ···················· ···················· ···················· ···················· ···················· ····················
W 3897 hCoV-19/United Arab Emirates/0733/2020|EPI_ISL_698 ···················· ···················· ···················· ···················· ···················· ····················
W 3898 hCoV-19/United Arab Emirates/0753/2020|EPI_ISL_698 ···················· ···················· ···················· ···················· ···················· ····················
W 3899 hCoV-19/United Arab Emirates/0788/2020|EPI_ISL_698 ···················· ···················· ···················· ···················· ···················· ····················
W 3900 hCoV-19/United Arab Emirates/0100/2020|EPI_ISL_698 ···················· ···················· ···················· ···················· ···················· ····················
W 3901 hCoV-19/United Arab Emirates/0491/2020|EPI_ISL_698 ···················· ···················· ···················· ···················· ···················· ····················
W 3902 hCoV-19/United Arab Emirates/0506/2020|EPI_ISL_698 ···················· ···················· ···················· ···················· ···················· ····················
W 3903 hCoV-19/United Arab Emirates/0513/2020|EPI_ISL_698 ···················· ···················· ···················· ···················· ···················· ····················
W 3904 hCoV-19/United Arab Emirates/0527/2020|EPI_ISL_698 ···················· ···················· ···················· ···················· ···················· ····················
W 3905 hCoV-19/United Arab Emirates/0736/2020|EPI_ISL_698 ···················· ···················· ···················· ···················· ···················· ····················
W 3906 hCoV-19/United Arab Emirates/0802/2020|EPI_ISL_698 ···················· ···················· ···················· ···················· ···················· ····················
W 3907 hCoV-19/United Arab Emirates/0971/2020|EPI_ISL_698 ···················· ···················· ···················· ···················· ···················· ····················
W 3908 hCoV-19/United Arab Emirates/0273/2020|EPI_ISL_698 ···················· ···················· ···················· ···················· ···················· ····················
W 3909 hCoV-19/United Arab Emirates/0133/2020|EPI_ISL_698 ···················· ···················· ···················· ···················· ···················· ····················
W 3910 hCoV-19/United Arab Emirates/0782/2020|EPI_ISL_698 ···················· ···················· ···················· ···················· ···················· ····················
W 3911 hCoV-19/United Arab Emirates/0480/2020|EPI_ISL_698 ···················· ···················· ···················· ···················· ···················· ····················
W 3912 hCoV-19/United Arab Emirates/0514/2020|EPI_ISL_698 ···················· ···················· ···················· ···················· ···················· ····················
W 3913 hCoV-19/United Arab Emirates/0560/2020|EPI_ISL_698 ···················· ···················· ···················· ···················· ···················· ····················
W 3914 hCoV-19/United Arab Emirates/0606/2020|EPI_ISL_698 ···················· ···················· ···················· ···················· ···················· ····················
W 3915 hCoV-19/United Arab Emirates/0572/2020|EPI_ISL_698 ···················· ···················· ···················· ···················· ···················· ····················
W 3916 hCoV-19/England/ALDP-BB5166/2020|EPI_ISL_702000|20 ···················· ·········T·········· ···················· ···················· ···················· ····················
W 3917 hCoV-19/England/ALDP-BB35C6/2020|EPI_ISL_702001|20 ···················· ···················· ···················· ···················· ···················· ····················
W 3918 hCoV-19/England/ALDP-BB3827/2020|EPI_ISL_702002|20 ···················· ···················· ···················· ···················· ···················· ····················
W 3919 hCoV-19/England/ALDP-BB3D91/2020|EPI_ISL_702003|20 ···················· ···················· ···················· ···················· ···················· ····················
W 3920 hCoV-19/England/ALDP-BB38DC/2020|EPI_ISL_702006|20 ···················· ···················· ···················· ···················· ···················· ····················
W 3921 hCoV-19/England/ALDP-BB50F0/2020|EPI_ISL_702007|20 ···················· ···················· ···················· ···················· ···················· ····················
W 3922 hCoV-19/England/ALDP-BB5403/2020|EPI_ISL_702008|20 ···················· ···················· ···················· ···················· ···················· ····················
W 3923 hCoV-19/England/ALDP-BBB160/2020|EPI_ISL_702009|20 ···················· ···················· ···················· ···················· ···················· ····················
W 3924 hCoV-19/England/QEUH-BC452C/2020|EPI_ISL_703300|20 ···················· ·A·················· ···················· ···················· ···················· ····················
W 3925 hCoV-19/England/ALDP-BCA5CC/2020|EPI_ISL_703301|20 ···················· ···················· ···················· ···················· ···················· ····················
W 3926 hCoV-19/England/QEUH-BC3F89/2020|EPI_ISL_703304|20 ···················· ···················· ···················· ···················· ···················· ····················
W 3927 hCoV-19/England/ALDP-BCA201/2020|EPI_ISL_703306|20 ···················· ···················· ···················· ···················· ···················· ····················
W 3928 hCoV-19/England/ALDP-BCAB9D/2020|EPI_ISL_703307|20 ···················· ···················· ···················· ···················· ···················· ····················
W 3929 hCoV-19/England/QEUH-BC3F3E/2020|EPI_ISL_703309|20 ···················· ···················· ···················· ···················· ···················· ····················
W 3930 hCoV-19/England/QEUH-BC44B6/2020|EPI_ISL_703320|20 ···················· ···················· ···················· ···················· ···················· ····················
W 3931 hCoV-19/England/ALDP-BCA429/2020|EPI_ISL_703322|20 ···················· ···················· ···················· ···················· ···················· ····················
W 3932 hCoV-19/England/QEUH-BC4744/2020|EPI_ISL_703323|20 ···················· ···················· ···················· ···················· ···················· ····················
W 3933 hCoV-19/England/QEUH-BC46B0/2020|EPI_ISL_703325|20 ···················· ···················· ···················· ···················· ···················· ····················
W 3934 hCoV-19/England/ALDP-BCA80F/2020|EPI_ISL_703326|20 ···················· ···················· ···················· ···················· ···················· ····················
W 3935 hCoV-19/England/ALDP-BCA140/2020|EPI_ISL_703328|20 ···················· ···················· ···················· ···················· ···················· ····················
W 3936 hCoV-19/England/ALDP-BCA4CF/2020|EPI_ISL_703329|20 ···················· ···················· ···················· ···················· ···················· ···A················
W 3937 hCoV-19/England/QEUH-BC4850/2020|EPI_ISL_703331|20 ···················· ···················· ···················· ···················· ···················· ····················
W 3938 hCoV-19/England/ALDP-BCA5AE/2020|EPI_ISL_703333|20 ···················· ···················· ···················· ···················· ···················· ····················
W 3939 hCoV-19/England/ALDP-BCA7D5/2020|EPI_ISL_703336|20 ···················· ···················· ···················· ···················· ···················· ····················
W 3940 hCoV-19/England/QEUH-BC45FF/2020|EPI_ISL_703337|20 ···················· ···················· ···················· ···················· ···················· ····················
W 3941 hCoV-19/England/ALDP-BCA91B/2020|EPI_ISL_703339|20 ···················· ···················· ···················· ···················· ···················· ····················
W 3942 hCoV-19/England/QEUH-BCD471/2020|EPI_ISL_704652|20 ···················· ···················· ···················· ···················· ······T············· ····················
W 3943 hCoV-19/England/QEUH-BCCB31/2020|EPI_ISL_704655|20 ···················· ···················· ···················· ···················· ···················· ····················
W 3944 hCoV-19/England/QEUH-BCD778/2020|EPI_ISL_704657|20 ···················· ···················· ···················· ···················· ···················· ····················
W 3945 hCoV-19/England/QEUH-BCD480/2020|EPI_ISL_704658|20 ···················· ···················· ···················· ···················· ···················· ····················
W 3946 hCoV-19/England/QEUH-BCB72F/2020|EPI_ISL_704663|20 ···················· ···················· ···················· ···················· ···················· ····················
W 3947 hCoV-19/England/QEUH-BCB75C/2020|EPI_ISL_704665|20 ···················· ···················· ···················· ···················· ···················· ····················
W 3948 hCoV-19/England/QEUH-BCB798/2020|EPI_ISL_704667|20 ···················· ···················· ···················· ···················· ···················· ····················
W 3949 hCoV-19/England/LIVE-DC4690/2020|EPI_ISL_705901|20 ···················· ···················· ···················· ···················· ···················· ····················
W 3950 hCoV-19/England/LIVE-DC46AF/2020|EPI_ISL_705902|20 ···················· ···················· ···················· ···················· ···················· ····················
W 3951 hCoV-19/England/LIVE-DC46CD/2020|EPI_ISL_705903|20 ···················· ···················· ···················· ···················· ···················· ····················
W 3952 hCoV-19/England/LIVE-DC46DC/2020|EPI_ISL_705904|20 ···················· ···················· ···················· ···················· ···················· ····················
W 3953 hCoV-19/England/LIVE-DC478E/2020|EPI_ISL_705905|20 ···················· ···················· ···················· ···················· ···················· ····················
W 3954 hCoV-19/England/LIVE-DC48C7/2020|EPI_ISL_705906|20 ···················· ···················· ···················· ···················· ···················· ····················
W 3955 hCoV-19/England/LIVE-DC48D6/2020|EPI_ISL_705907|20 ···················· ···················· ···················· ···················· ···················· ····················
W 3956 hCoV-19/England/LIVE-DC4900/2020|EPI_ISL_705908|20 ···················· ···················· ···················· ···················· ···················· ····················
W 3957 hCoV-19/England/LIVE-DC4988/2020|EPI_ISL_705909|20 ···················· ·········T·········· ···················· ···················· ···················· ····················
W 3958 hCoV-19/England/LIVE-DC4A2B/2020|EPI_ISL_705910|20 ···················· ···················· ···················· ···················· ···················· ····················
W 3959 hCoV-19/England/LIVE-DC4A49/2020|EPI_ISL_705911|20 ···················· ···················· ···················· ···················· ···················· ····················
W 3960 hCoV-19/England/LIVE-DC4A85/2020|EPI_ISL_705912|20 ···················· ···················· ···················· ···················· ···················· ····················

W 3961 hCoV-19/England/LIVE-DC4C8F/2020|EPI_ISL_705913|20 ···················· ···················· ····················· ···················· ···················· ····················
W 3962 hCoV-19/England/LIVE-DC4C9E/2020|EPI_ISL_705914|20 ···················· ···················· ····················· ···················· ···················· ····················
W 3963 hCoV-19/England/LIVE-DC4CCB/2020|EPI_ISL_705915|20 ···················· ···················· ····················· ···················· ···················· ····················
W 3964 hCoV-19/England/LIVE-DC4D8C/2020|EPI_ISL_705916|20 ···················· ···················· ····················· ···················· ···················· ····················
W 3965 hCoV-19/England/LIVE-DC4DD7/2020|EPI_ISL_705917|20 ···················· ···················· ····················· ···················· ···················· ····················
W 3966 hCoV-19/England/LIVE-DC4DF5/2020|EPI_ISL_705918|20 ···················· ···················· ····················· ···················· ···················· ····················
W 3967 hCoV-19/England/LIVE-DC4E5C/2020|EPI_ISL_705919|20 ···················· ···················· ····················· ···················· ···················· ····················
W 3968 hCoV-19/England/LIVE-DC4ED4/2020|EPI_ISL_705920|20 ···················· ···················· ····················· ···················· ···················· ····················
W 3969 hCoV-19/England/LIVE-DC65EC/2020|EPI_ISL_705921|20 ···················· ···················· ····················· ···················· ···················· ····················
W 3970 hCoV-19/England/LIVE-DC677D/2020|EPI_ISL_705922|20 ···················· ···················· ····················· ···················· ···················· ····················
W 3971 hCoV-19/England/LIVE-DC6A1A/2020|EPI_ISL_705923|20 ···················· ···················· ····················· ···················· ···················· ····················
W 3972 hCoV-19/England/LIVE-DC6CBA/2020|EPI_ISL_705924|20 ···················· ···················· ····················· ···················· ···················· ····················
W 3973 hCoV-19/England/LIVE-DC6E2D/2020|EPI_ISL_705925|20 ···················· ···················· ····················· ···················· ···················· ····················
W 3974 hCoV-19/England/LIVE-DC6E3C/2020|EPI_ISL_705926|20 ···················· ···················· ····················· ···················· ···················· ····················
W 3977 hCoV-19/England/LIVE-DC7D3E/2020|EPI_ISL_705929|20 ···················· ···················· ····················· ···················· ···················· ····················
W 3978 hCoV-19/England/LIVE-DC7D4D/2020|EPI_ISL_705930|20 ···················· ···················· ····················· ···················· ···················· ····················
W 3979 hCoV-19/England/LIVE-DC7E2C/2020|EPI_ISL_705931|20 ···················· ···················· ····················· ···················· ···················· ····················
W 3980 hCoV-19/England/LIVE-DC7E3B/2020|EPI_ISL_705932|20 ···················· ···················· ····················· ···················· ···················· ····················
W 3981 hCoV-19/England/LIVE-DC7E4A/2020|EPI_ISL_705933|20 ···················· ···················· ····················· ···················· ···················· ····················
W 3982 hCoV-19/England/LIVE-DC7E59/2020|EPI_ISL_705934|20 ···················· ···················· ····················· ···················· ···················· ····················
W 3983 hCoV-19/England/LIVE-DC80F8/2020|EPI_ISL_705935|20 ···················· ···················· ····················· ···················· ···················· ····················
W 3984 hCoV-19/England/LIVE-DC8131/2020|EPI_ISL_705936|20 ···················· ···················· ····················· ···················· ···················· ····················
W 3985 hCoV-19/England/LIVE-DC81F5/2020|EPI_ISL_705937|20 ···················· ···················· ····················· ···················· ···················· ····················
W 3986 hCoV-19/England/LIVE-DC824D/2020|EPI_ISL_705938|20 ···················· ···················· ····················· ···················· ···················· ····················
W 3987 hCoV-19/England/LIVE-DC825C/2020|EPI_ISL_705939|20 ···················· ···················· ····················· ···················· ···················· ····················
W 3988 hCoV-19/England/LIVE-DC8B33/2020|EPI_ISL_705940|20 ···················· ···················· ····················· ···················· ···················· ····················
W 3989 hCoV-19/England/LIVE-DC9BC9/2020|EPI_ISL_705941|20 ···················· ···················· ····················· ···················· ···················· ····················
W 3990 hCoV-19/England/LOND-1251A67/2020|EPI_ISL_705942|2 ···················· ···················· ····················· ···················· ···················· ····················
W 3991 hCoV-19/England/LOND-1251A76/2020|EPI_ISL_705943|2 ···················· ···················· ····················· ···················· ·····T·············· ····················
W 3992 hCoV-19/England/LOND-1251A94/2020|EPI_ISL_705944|2 ···················· ···················· ····················· ···················· ···················· ····················
W 3994 hCoV-19/England/LOND-1251AC1/2020|EPI_ISL_705946|2 ···················· ···················· ····················· ···················· ···················· ····················
W 3995 hCoV-19/England/LOND-1251AEF/2020|EPI_ISL_705947|2 ···················· ···················· ····················· ···················· ···················· ····················
W 3996 hCoV-19/England/LOND-1251B0A/2020|EPI_ISL_705948|2 ···················· ···················· ····················· ···················· ···················· ····················
W 3997 hCoV-19/England/LOND-1251B37/2020|EPI_ISL_705949|2 ···················· ···················· ····················· ···················· ···················· ····················
W 3998 hCoV-19/England/LOND-1251B55/2020|EPI_ISL_705950|2 ···················· ···················· ····················· ···················· ···················· ····················
W 4000 hCoV-19/England/LOND-1251B91/2020|EPI_ISL_705952|2 ···················· ···················· ····················· ···················· ···················· ····················
W 4001 hCoV-19/England/LOND-1251BBF/2020|EPI_ISL_705953|2 ···················· ···················· ····················· ···················· ···················· ····················
W 4002 hCoV-19/England/LOND-1251BFB/2020|EPI_ISL_705954|2 ···················· ···················· ····················· ···················· ···················· ····················
W 4003 hCoV-19/England/LOND-1251CBC/2020|EPI_ISL_705955|2 ···················· ···················· ····················· ···················· ···················· ····················
W 4005 hCoV-19/England/LOND-1251F0E/2020|EPI_ISL_705957|2 ···················· ···················· ····················· ···················· ···················· ····················
W 4006 hCoV-19/England/LOND-1251F2C/2020|EPI_ISL_705958|2 ···················· ···················· ····················· ···················· ···················· ····················
W 4007 hCoV-19/England/LOND-1251F4A/2020|EPI_ISL_705959|2 ···················· ···················· ····················· ···················· ···················· ····················
W 4009 hCoV-19/England/LOND-1264278/2020|EPI_ISL_705961|2 ···················· ···················· ····················· ···················· ···················· ····················
W 4010 hCoV-19/England/LOND-1264357/2020|EPI_ISL_705962|2 ···················· ···················· ····················· ···················· ···················· ····················
W 4011 hCoV-19/England/LOND-12643C0/2020|EPI_ISL_705963|2 ···················· ···················· ····················· ···················· ···················· ····················
W 4012 hCoV-19/England/LOND-1264481/2020|EPI_ISL_705964|2 ···················· ···················· ····················· ···················· ···················· ····················
W 4013 hCoV-19/England/LOND-12644FA/2020|EPI_ISL_705965|2 ···················· ···················· ····················· ···················· ···················· ····················
W 4014 hCoV-19/England/LOND-126458E/2020|EPI_ISL_705966|2 ···················· ···················· ····················· ···················· ···················· ····················
W 4015 hCoV-19/England/LOND-126459D/2020|EPI_ISL_705967|2 ···················· ···················· ····················· ···················· ···················· ····················
W 4016 hCoV-19/England/LOND-1264630/2020|EPI_ISL_705968|2 ···················· ···················· ····················· ···················· ···················· ····················
W 4017 hCoV-19/England/LOND-12646B8/2020|EPI_ISL_705969|2 ···················· ···················· ····················· ···················· ···················· ····················
W 4018 hCoV-19/England/MILK-A91644/2020|EPI_ISL_705972|20 ···················· ···················· ····················· ···················· ···················· ····················
W 4019 hCoV-19/England/MILK-A91662/2020|EPI_ISL_705973|20 ···················· ···················· ····················· ···················· ···················· ····················
W 4020 hCoV-19/England/MILK-A91732/2020|EPI_ISL_705980|20 ···················· ···················· ····················· ···················· ···················· ····················
W 4021 hCoV-19/England/MILK-A91741/2020|EPI_ISL_705981|20 ···················· ···················· ····················· ···················· ···················· ····················
W 4022 hCoV-19/England/MILK-A9177E/2020|EPI_ISL_705982|20 ···················· ···················· ····················· ···················· ···················· ····················
W 4023 hCoV-19/England/MILK-A9178D/2020|EPI_ISL_705983|20 ···················· ···················· ····················· ···················· ···················· ····················
W 4024 hCoV-19/England/MILK-A9179C/2020|EPI_ISL_705984|20 ···················· ···················· ····················· ···················· ···················· ····················
W 4025 hCoV-19/England/MILK-A917AB/2020|EPI_ISL_705985|20 ···················· ···················· ····················· ···················· ···················· ····················
W 4026 hCoV-19/England/MILK-A917BA/2020|EPI_ISL_705986|20 ···················· ···················· ····················· ···················· ···················· ····················
W 4027 hCoV-19/England/MILK-A917D8/2020|EPI_ISL_705987|20 ···················· ·A·················· ····················· ···················· ···················· ··A·················
W 4028 hCoV-19/England/MILK-A917F6/2020|EPI_ISL_705988|20 ···················· ···················· ····················· ···················· ···················· ····················
W 4029 hCoV-19/England/MILK-A91811/2020|EPI_ISL_705989|20 ···················· ···················· ····················· ···················· ···················· ····················
W 4030 hCoV-19/England/MILK-A91820/2020|EPI_ISL_705990|20 ···················· ···················· ····················· ···················· ···················· ····················
W 4031 hCoV-19/England/MILK-A9183F/2020|EPI_ISL_705991|20 ···················· ···················· ····················· ···················· ···················· ····················
W 4032 hCoV-19/England/MILK-A9185D/2020|EPI_ISL_705992|20 ···················· ···················· ····················· ···················· ···················· ····················
W 4033 hCoV-19/England/MILK-A9186C/2020|EPI_ISL_705993|20 ···················· ···················· ····················· ···················· ···················· ····················
W 4034 hCoV-19/England/MILK-A918B7/2020|EPI_ISL_705994|20 ···················· ···················· ····················· ···················· ···················· ····················
W 4035 hCoV-19/England/MILK-A918E4/2020|EPI_ISL_705995|20 ···················· ···················· ····················· ···················· ···················· ····················
W 4036 hCoV-19/England/MILK-A918F3/2020|EPI_ISL_705996|20 ···················· ···················· ····················· ···················· ···················· ····················
W 4037 hCoV-19/England/MILK-A9195A/2020|EPI_ISL_705997|20 ···················· ···················· ····················· ···················· ···················· ····················
W 4038 hCoV-19/England/MILK-A91987/2020|EPI_ISL_705998|20 ···················· ···················· ····················· ···················· ···················· ····················
W 4039 hCoV-19/England/MILK-A919B4/2020|EPI_ISL_705999|20 ···················· ···················· ····················· ···················· ···················· ····················
W 4040 hCoV-19/Norway/5713/2020|EPI_ISL_708150|2020-11-11 ···················· ·A·················· ····················· ···················· ···················· ··A·················
W 4041 hCoV-19/Norway/5732/2020|EPI_ISL_708151|2020-11-14 ···················· ·A·················· ····················· ···················· ···················· ··A·················
W 4042 hCoV-19/Norway/5733/2020|EPI_ISL_708152|2020-11-12 ···················· ···················· ····················· ···················· ···················· ····················
W 4043 hCoV-19/Norway/5735/2020|EPI_ISL_708153|2020-11-13 ···················· ·A·················· ····················· ···················· ···················· ··A·················
W 4044 hCoV-19/Norway/5738/2020|EPI_ISL_708154|2020-10-27 ···················· ···················· ····················· ···················· ···················· ····················
W 4045 hCoV-19/Norway/5739/2020|EPI_ISL_708155|2020-10-27 ···················· ···················· ····················· ···················· ···················· ····················
W 4046 hCoV-19/Norway/5745/2020|EPI_ISL_708156|2020-10-28 ···················· ···················· ····················· ···················· ···················· ····················

| | | | | | | | |
|---|---|---|---|---|---|---|---|
| W | 4047 hCoV-19/Norway/5746/2020\|EPI_ISL_708157\|2020-10-26 | ···················· | ···················· | ····················· | ···················· | ···················· | ···················· |
| W | 4048 hCoV-19/Norway/5749/2020\|EPI_ISL_708158\|2020-10-29 | ···················· | ···················· | ····················· | ···················· | ···················· | ···················· |
| W | 4049 hCoV-19/Norway/5750/2020\|EPI_ISL_708159\|2020-10-29 | ···················· | ·A·················· | ····················· | ···················· | ···················· | ···A················ |
| W | 4050 hCoV-19/Norway/5753/2020\|EPI_ISL_708160\|2020-10-29 | ···················· | ·A·················· | ····················· | ···················· | ···················· | ···A················ |
| W | 4051 hCoV-19/Norway/5755/2020\|EPI_ISL_708161\|2020-10-28 | ···················· | ···················· | ····················· | ···················· | ···················· | ···················· |
| W | 4052 hCoV-19/Norway/5756/2020\|EPI_ISL_708162\|2020-10-28 | ···················· | ···················· | ····················· | ···················· | ···················· | ···················· |
| W | 4053 hCoV-19/Norway/5757/2020\|EPI_ISL_708163\|2020-10-19 | ···················· | ···················· | ····················· | ···················· | ···················· | ···················· |
| W | 4054 hCoV-19/Norway/5761/2020\|EPI_ISL_708164\|2020-10-24 | ···················· | ···················· | ····················· | ···················· | ···················· | ···················· |
| W | 4055 hCoV-19/Norway/5762/2020\|EPI_ISL_708165\|2020-10-26 | ···················· | ···················· | ····················· | ···················· | ···················· | ···················· |
| W | 4056 hCoV-19/Norway/5764/2020\|EPI_ISL_708166\|2020-10-26 | ···················· | ···················· | ····················· | ···················· | ···················· | ···················· |
| W | 4057 hCoV-19/Norway/5766/2020\|EPI_ISL_708167\|2020-10-27 | ···················· | ···················· | ····················· | ···················· | ···················· | ···················· |
| W | 4058 hCoV-19/Norway/5767/2020\|EPI_ISL_708168\|2020-10-27 | ···················· | ···················· | ····················· | ···················· | ···················· | ···················· |
| W | 4059 hCoV-19/Norway/5777/2020\|EPI_ISL_708169\|2020-11-18 | ···················· | ···················· | ····················· | ···················· | ···················· | ···················· |
| W | 4060 hCoV-19/Norway/5810/2020\|EPI_ISL_708180\|2020-11-17 | ···················· | ·A·················· | ····················· | ···················· | ···················· | ···A················ |
| W | 4061 hCoV-19/Norway/5812/2020\|EPI_ISL_708181\|2020-11-16 | ···················· | ···················· | ····················· | ···················· | ···················· | ···················· |
| W | 4062 hCoV-19/USA/CA-LACPHL-AD00007/2020\|EPI_ISL_708182\| | ···················· | ···················· | ····················· | ···················· | ···················· | ···················· |
| W | 4063 hCoV-19/USA/CA-LACPHL-AD00009/2020\|EPI_ISL_708183\| | ···················· | ···················· | ····················· | ···················· | ···················· | ···················· |
| W | 4064 hCoV-19/Turkey/DEN-PAU03/2020\|EPI_ISL_708184\|2020- | ···················· | ···················· | ····················· | ···················· | ···················· | ···················· |
| W | 4065 hCoV-19/Turkey/DEN-PAU04/2020\|EPI_ISL_708185\|2020- | ···················· | ···················· | ····················· | ···················· | ···················· | ···················· |
| W | 4066 hCoV-19/Turkey/DEN-PAU08/2020\|EPI_ISL_708186\|2020- | ···················· | ···················· | ····················· | ···················· | ···················· | ···················· |
| W | 4067 hCoV-19/Turkey/DEN-PAU10/2020\|EPI_ISL_708188\|2020- | ···················· | ···················· | ····················· | ···················· | ···················· | ···················· |
| W | 4068 hCoV-19/Turkey/DEN-PAU11/2020\|EPI_ISL_708189\|2020- | ···················· | ···················· | ····················· | ···················· | ···················· | ···················· |
| W | 4069 hCoV-19/Turkey/DEN-PAU12/2020\|EPI_ISL_708190\|2020- | ···················· | ···················· | ····················· | ···················· | ···················· | ···················· |
| W | 4070 hCoV-19/Turkey/DEN-PAU15/2020\|EPI_ISL_708192\|2020- | ···················· | ···················· | ····················· | ···················· | ···················· | ···················· |
| W | 4071 hCoV-19/Turkey/DEN-PAU17/2020\|EPI_ISL_708193\|2020- | ···················· | ····G··············· | ····················· | ···················· | ···················· | ···················· |
| W | 4072 hCoV-19/Turkey/DEN-PAU19/2020\|EPI_ISL_708194\|2020- | ···················· | ···················· | ····················· | ···················· | ···················· | ···················· |
| W | 4073 hCoV-19/Turkey/DEN-PAU14/2020\|EPI_ISL_708195\|2020- | ···················· | ···················· | ····················· | ···················· | ···················· | ···················· |
| W | 4074 hCoV-19/Norway/5116/2020\|EPI_ISL_708196\|2020-10-29 | ···················· | ····G··············· | ····················· | ···················· | ···················· | ···················· |
| W | 4075 hCoV-19/Norway/5382/2020\|EPI_ISL_708197\|2020-10-27 | ···················· | ···················· | ····················· | ···················· | ···················· | ···················· |
| W | 4076 hCoV-19/Norway/5400/2020\|EPI_ISL_708198\|2020-10-28 | ···················· | ···················· | ····················· | ···················· | ···················· | ···················· |
| W | 4077 hCoV-19/Norway/5542/2020\|EPI_ISL_708199\|2020-11-07 | ···················· | ····G··············· | ····················· | ···················· | ···················· | ···················· |
| W | 4078 hCoV-19/England/ALDP-C067DC/2020\|EPI_ISL_709480\|20 | ···················· | ···················· | ····················· | ···················· | ···················· | ···················· |
| W | 4079 hCoV-19/England/QEUH-C1600B/2020\|EPI_ISL_709483\|20 | ···················· | ···················· | ····················· | ···················· | ···················· | ···················· |
| W | 4080 hCoV-19/England/ALDP-C069B8/2020\|EPI_ISL_709484\|20 | ···················· | ···················· | ····················· | ···················· | ···················· | ···················· |
| W | 4081 hCoV-19/England/QEUH-C14C45/2020\|EPI_ISL_709486\|20 | ···················· | ···················· | ····················· | ···················· | ···················· | ···················· |
| W | 4082 hCoV-19/England/QEUH-C16065/2020\|EPI_ISL_709487\|20 | ···················· | ···················· | ····················· | ···················· | ···················· | ···················· |
| W | 4083 hCoV-19/England/ALDP-C05F7B/2020\|EPI_ISL_709488\|20 | ···················· | ···················· | ····················· | ···················· | ···················· | ···················· |
| W | 4084 hCoV-19/England/QEUH-C15FC3/2020\|EPI_ISL_709490\|20 | ···················· | ···················· | ····················· | ···················· | ···················· | ···················· |
| W | 4085 hCoV-19/England/ALDP-C061B0/2020\|EPI_ISL_709491\|20 | ···················· | ····A··············· | ····················· | ···················· | ···················· | ···················· |
| W | 4086 hCoV-19/England/ALDP-C06E04/2020\|EPI_ISL_709492\|20 | ···················· | ···················· | ····················· | ···················· | ···················· | ···················· |
| W | 4087 hCoV-19/England/QEUH-C149A8/2020\|EPI_ISL_709494\|20 | ···················· | ···················· | ····················· | ···················· | ···················· | ···················· |
| W | 4088 hCoV-19/England/QEUH-C14586/2020\|EPI_ISL_709496\|20 | ···················· | ···················· | ····················· | ···················· | ···················· | ···················· |
| W | 4089 hCoV-19/England/QEUH-C159D4/2020\|EPI_ISL_709497\|20 | ···················· | ···················· | ····················· | ···················· | ···················· | ···················· |
| W | 4090 hCoV-19/England/QEUH-C14692/2020\|EPI_ISL_709498\|20 | ···················· | ···················· | ····················· | ···················· | ···················· | ···················· |
| W | 4091 hCoV-19/Palestine/AAS15/2020\|EPI_ISL_710483\|2020-1 | ···················· | ···················· | ····················· | ···················· | ···················· | ···················· |
| W | 4092 hCoV-19/Palestine/AAS16/2020\|EPI_ISL_710484\|2020-1 | ···················· | ···················· | ····················· | ···················· | ···················· | ···················· |
| W | 4093 hCoV-19/Spain/AS-HUCA-232017735/2020\|EPI_ISL_71048 | ···················· | ···················· | ····················· | ···················· | ···················· | ···················· |
| W | 4094 hCoV-19/Spain/AS-HUCA-232015439/2020\|EPI_ISL_71048 | ···················· | ···················· | ····················· | ···················· | ···················· | ···················· |
| W | 4095 hCoV-19/Spain/AS-HUCA-232026126/2020\|EPI_ISL_71048 | ···················· | ···················· | ····················· | ···················· | ···················· | ···················· |
| W | 4096 hCoV-19/Spain/AS-HUCA-232023718/2020\|EPI_ISL_71049 | ···················· | ···················· | ····················· | ···················· | ···················· | ···················· |
| W | 4097 hCoV-19/USA/AZ-TG371263/2020\|EPI_ISL_694523\|2020-0 | ···················· | ···················· | ····················· | ···················· | ···················· | ···················· |
| W | 4098 hCoV-19/USA/AZ-TG371271/2020\|EPI_ISL_694530\|2020-0 | ···················· | ···················· | ····················· | ···················· | ···················· | ···················· |
| W | 4099 hCoV-19/USA/AZ-TG371272/2020\|EPI_ISL_694531\|2020-0 | ···················· | ···················· | ····················· | ···················· | ···················· | ···················· |
| W | 4100 hCoV-19/USA/AZ-TG371274/2020\|EPI_ISL_694533\|2020-0 | ···················· | ···················· | ····················· | ···················· | ···················· | ···················· |
| W | 4101 hCoV-19/USA/AZ-TG371280/2020\|EPI_ISL_694539\|2020-0 | ···················· | ···················· | ····················· | ···················· | ···················· | ···················· |
| W | 4102 hCoV-19/USA/AZ-TG371282/2020\|EPI_ISL_694541\|2020-0 | ···················· | ···················· | ····················· | ···················· | ···················· | ···················· |
| W | 4103 hCoV-19/USA/AZ-TG371283/2020\|EPI_ISL_694542\|2020-0 | ···················· | ···················· | ····················· | ···················· | ···················· | ···················· |
| W | 4104 hCoV-19/USA/AZ-TG371284/2020\|EPI_ISL_694543\|2020-0 | ···················· | ···················· | ····················· | ···················· | ···················· | ···················· |
| W | 4105 hCoV-19/USA/AZ-TG371286/2020\|EPI_ISL_694545\|2020-0 | ···················· | ···················· | ····················· | ···················· | ···················· | ···················· |
| W | 4106 hCoV-19/USA/AZ-TG371288/2020\|EPI_ISL_694547\|2020-0 | ···················· | ···················· | ····················· | ···················· | ···················· | ···················· |
| W | 4107 hCoV-19/USA/AZ-TG371289/2020\|EPI_ISL_694548\|2020-0 | ···················· | ···················· | ····················· | ···················· | ···················· | ···················· |
| W | 4108 hCoV-19/USA/AZ-TG371291/2020\|EPI_ISL_694550\|2020-0 | ···················· | ···················· | ····················· | ···················· | ···················· | ···················· |
| W | 4109 hCoV-19/USA/AZ-TG371295/2020\|EPI_ISL_694551\|2020-0 | ···················· | ···················· | ····················· | ···················· | ···················· | ···················· |
| W | 4110 hCoV-19/USA/AZ-TG371296/2020\|EPI_ISL_694552\|2020-0 | ···················· | ·A·················· | ····················· | ···················· | ···················· | ···················· |
| W | 4111 hCoV-19/USA/AZ-TG371297/2020\|EPI_ISL_694553\|2020-0 | ···················· | ···················· | ····················· | ···················· | ···················· | ···················· |
| W | 4112 hCoV-19/USA/AZ-TG371298/2020\|EPI_ISL_694554\|2020-0 | ···················· | ···················· | ····················· | ···················· | ···················· | ···················· |
| W | 4113 hCoV-19/USA/AZ-TG371301/2020\|EPI_ISL_694556\|2020-0 | ···················· | ···················· | ····················· | ···················· | ···················· | ···················· |
| W | 4114 hCoV-19/USA/AZ-TG371303/2020\|EPI_ISL_694558\|2020-0 | ···················· | ···················· | ····················· | ···················· | ···················· | ···················· |
| W | 4115 hCoV-19/USA/AZ-TG371442/2020\|EPI_ISL_694599\|2020-0 | ···················· | ···················· | ····················· | ···················· | ···················· | ···················· |
| W | 4116 hCoV-19/USA/AZ-TG648590/2020\|EPI_ISL_695842\|2020-0 | ···················· | ···················· | ····················· | ···················· | ···················· | ···················· |
| W | 4117 hCoV-19/USA/AZ-TG648593/2020\|EPI_ISL_695845\|2020-0 | ···················· | ···················· | ····················· | ···················· | ···················· | ···················· |
| W | 4118 hCoV-19/USA/AZ-TG648597/2020\|EPI_ISL_695848\|2020-0 | ···················· | ···················· | ····················· | ···················· | ···················· | ···················· |
| W | 4119 hCoV-19/USA/AZ-TG648601/2020\|EPI_ISL_695851\|2020-0 | ···················· | ···················· | ····················· | ···················· | ···················· | ···················· |
| W | 4120 hCoV-19/USA/AZ-TG648603/2020\|EPI_ISL_695852\|2020-0 | ···················· | ···················· | ····················· | ···················· | ···················· | ···················· |
| W | 4121 hCoV-19/USA/AZ-TG648604/2020\|EPI_ISL_695853\|2020-0 | ···················· | ····A··············· | ····················· | ···················· | ···················· | ···················· |
| W | 4122 hCoV-19/USA/AZ-TG648607/2020\|EPI_ISL_695854\|2020-0 | ···················· | ···················· | ····················· | ···················· | ···················· | ···················· |
| W | 4123 hCoV-19/USA/AZ-TG648616/2020\|EPI_ISL_695859\|2020-0 | ···················· | ···················· | ····················· | ···················· | ···················· | ···················· |
| W | 4124 hCoV-19/USA/AZ-TG648628/2020\|EPI_ISL_695865\|2020-0 | ···················· | ···················· | ····················· | ···················· | ···················· | ···················· |
| W | 4125 hCoV-19/USA/AZ-TG648636/2020\|EPI_ISL_695872\|2020-0 | ···················· | ···················· | ····················· | ···················· | ···················· | ···················· |
| W | 4126 hCoV-19/USA/AZ-TG648638/2020\|EPI_ISL_695873\|2020-0 | ···················· | ···················· | ····················· | ···················· | ···················· | ···················· |

| | | | | | | | | |
|---|---|---|---|---|---|---|---|---|
| W | 4127 | hCoV-19/USA/AZ-TG648640/2020\|EPI_ISL_695875\|2020-0 | ···················· | ···················· | ···················· | ···················· | ···················· | ···················· |
| W | 4128 | hCoV-19/USA/AZ-TG648647/2020\|EPI_ISL_695879\|2020-0 | ···················· | ···················· | ···················· | ···················· | ···················· | ···················· |
| W | 4129 | hCoV-19/USA/AZ-TG648649/2020\|EPI_ISL_695880\|2020-0 | ···················· | ···················· | ···················· | ···················· | ···················· | ···················· |
| W | 4130 | hCoV-19/USA/AZ-TG648653/2020\|EPI_ISL_695881\|2020-0 | ···················· | ···················· | ···················· | ···················· | ···················· | ···················· |
| W | 4131 | hCoV-19/USA/AZ-TG648664/2020\|EPI_ISL_695885\|2020-0 | ···················· | ···················· | ···················· | ···················· | ···················· | ···················· |
| W | 4132 | hCoV-19/USA/AZ-TG648674/2020\|EPI_ISL_695889\|2020-0 | ···················· | ···················· | ···················· | ···················· | ···················· | ···················· |
| W | 4133 | hCoV-19/United Arab Emirates/1036/2020\|EPI_ISL_698 | ···················· | ···················· | ···················· | ···················· | ···················· | ···················· |
| W | 4134 | hCoV-19/United Arab Emirates/1037/2020\|EPI_ISL_698 | ···················· | ···················· | ···················· | ···················· | ···················· | ···················· |
| W | 4135 | hCoV-19/United Arab Emirates/1042/2020\|EPI_ISL_698 | ···················· | ···················· | ···················· | ···················· | ···················· | ···················· |
| W | 4136 | hCoV-19/United Arab Emirates/1043/2020\|EPI_ISL_698 | ···················· | ···················· | ···················· | ···················· | ···················· | ···················· |
| W | 4137 | hCoV-19/United Arab Emirates/1044/2020\|EPI_ISL_698 | ···················· | ···················· | ···················· | ···················· | ···················· | ···················· |
| W | 4138 | hCoV-19/United Arab Emirates/1045/2020\|EPI_ISL_698 | ···················· | ···················· | ···················· | ···················· | ···················· | ···················· |
| W | 4139 | hCoV-19/United Arab Emirates/1046/2020\|EPI_ISL_698 | ···················· | ···················· | ···················· | ···················· | ···················· | ···················· |
| W | 4140 | hCoV-19/United Arab Emirates/1048/2020\|EPI_ISL_698 | ···················· | ···················· | ···················· | ···················· | ···················· | ···················· |
| W | 4141 | hCoV-19/United Arab Emirates/1049/2020\|EPI_ISL_698 | ···················· | ···················· | ···················· | ···················· | ···················· | ···················· |
| W | 4142 | hCoV-19/United Arab Emirates/1050/2020\|EPI_ISL_698 | ···················· | ···················· | ···················· | ···················· | ···················· | ···················· |
| W | 4143 | hCoV-19/United Arab Emirates/1051/2020\|EPI_ISL_698 | ···················· | ···················· | ···················· | ···················· | ···················· | ···················· |
| W | 4144 | hCoV-19/United Arab Emirates/1052/2020\|EPI_ISL_698 | ···················· | ···················· | ···················· | ···················· | ···················· | ···················· |
| W | 4145 | hCoV-19/United Arab Emirates/1053/2020\|EPI_ISL_698 | ···················· | ···················· | ···················· | ···················· | ···················· | ···················· |
| W | 4146 | hCoV-19/United Arab Emirates/1054/2020\|EPI_ISL_698 | ···················· | ···················· | ···················· | ···················· | ···················· | ···················· |
| W | 4147 | hCoV-19/United Arab Emirates/1055/2020\|EPI_ISL_698 | ···················· | ···················· | ···················· | ···················· | ···················· | ···················· |
| W | 4148 | hCoV-19/United Arab Emirates/1057/2020\|EPI_ISL_698 | ···················· | ···················· | ···················· | ···················· | ···················· | ···················· |
| W | 4149 | hCoV-19/United Arab Emirates/1058/2020\|EPI_ISL_698 | ···················· | ···················· | ···················· | ···················· | ···················· | ···················· |
| W | 4150 | hCoV-19/United Arab Emirates/1059/2020\|EPI_ISL_698 | ···················· | ···················· | ···················· | ···················· | ···················· | ···················· |
| W | 4151 | hCoV-19/United Arab Emirates/1061/2020\|EPI_ISL_698 | ···················· | ···················· | ···················· | ···················· | ···················· | ···················· |
| W | 4152 | hCoV-19/United Arab Emirates/1062/2020\|EPI_ISL_698 | ···················· | ···················· | ···················· | ···················· | ···················· | ···················· |
| W | 4153 | hCoV-19/United Arab Emirates/1063/2020\|EPI_ISL_698 | ···················· | ···················· | ···················· | ···················· | ···················· | ···················· |
| W | 4154 | hCoV-19/United Arab Emirates/1064/2020\|EPI_ISL_698 | ···················· | ···················· | ···················· | ···················· | ···················· | ···················· |
| W | 4155 | hCoV-19/United Arab Emirates/1065/2020\|EPI_ISL_698 | ···················· | ···················· | ···················· | ···················· | ···················· | ···················· |
| W | 4156 | hCoV-19/United Arab Emirates/1066/2020\|EPI_ISL_698 | ···················· | ···················· | ···················· | ···················· | ···················· | ···················· |
| W | 4157 | hCoV-19/United Arab Emirates/1067/2020\|EPI_ISL_698 | ···················· | ···················· | ···················· | ···················· | ···················· | ···················· |
| W | 4158 | hCoV-19/United Arab Emirates/1068/2020\|EPI_ISL_698 | ···················· | ···················· | ···················· | ···················· | ···················· | ···················· |
| W | 4159 | hCoV-19/United Arab Emirates/1070/2020\|EPI_ISL_698 | ···················· | ···················· | ···················· | ···················· | ···················· | ···················· |
| W | 4160 | hCoV-19/United Arab Emirates/1071/2020\|EPI_ISL_698 | ···················· | ···················· | ···················· | ···················· | ···················· | ···················· |
| W | 4161 | hCoV-19/United Arab Emirates/1072/2020\|EPI_ISL_698 | ···················· | ···················· | ···················· | ···················· | ···················· | ···················· |
| W | 4162 | hCoV-19/United Arab Emirates/1073/2020\|EPI_ISL_698 | ···················· | ···················· | ···················· | ···················· | ···················· | ···················· |
| W | 4163 | hCoV-19/United Arab Emirates/1074/2020\|EPI_ISL_698 | ···················· | ···················· | ···················· | ···················· | ···················· | ···················· |
| W | 4164 | hCoV-19/United Arab Emirates/1075/2020\|EPI_ISL_698 | ···················· | ···················· | ···················· | ···················· | ···················· | ···················· |
| W | 4165 | hCoV-19/United Arab Emirates/1076/2020\|EPI_ISL_698 | ···················· | ···················· | ···················· | ···················· | ···················· | ···················· |
| W | 4166 | hCoV-19/United Arab Emirates/1077/2020\|EPI_ISL_698 | ···················· | ···················· | ···················· | ···················· | ···················· | ···················· |
| W | 4167 | hCoV-19/United Arab Emirates/1078/2020\|EPI_ISL_698 | ···················· | ···················· | ···················· | ···················· | ···················· | ···················· |
| W | 4168 | hCoV-19/United Arab Emirates/1079/2020\|EPI_ISL_698 | ···················· | ···················· | ···················· | ···················· | ···················· | ···················· |
| W | 4169 | hCoV-19/United Arab Emirates/1080/2020\|EPI_ISL_698 | ···················· | ···················· | ···················· | ···················· | ···················· | ···················· |
| W | 4170 | hCoV-19/United Arab Emirates/1081/2020\|EPI_ISL_698 | ···················· | ···················· | ···················· | ···················· | ···················· | ···················· |
| W | 4171 | hCoV-19/United Arab Emirates/1082/2020\|EPI_ISL_698 | ···················· | ···················· | ···················· | ···················· | ···················· | ···················· |
| W | 4172 | hCoV-19/United Arab Emirates/1083/2020\|EPI_ISL_698 | ···················· | ···················· | ···················· | ···················· | ···················· | ···················· |
| W | 4173 | hCoV-19/United Arab Emirates/1084/2020\|EPI_ISL_698 | ···················· | ···················· | ···················· | ···················· | ···················· | ···················· |
| W | 4174 | hCoV-19/United Arab Emirates/1085/2020\|EPI_ISL_698 | ···················· | ···················· | ···················· | ···················· | ···················· | ···················· |
| W | 4175 | hCoV-19/Thailand/Phuket_SQ_AR0013/2020\|EPI_ISL_699 | ···················· | ···················· | ···················· | ···················· | ···················· | ···················· |
| W | 4176 | hCoV-19/Thailand/Bangkok_PT386/2020\|EPI_ISL_699484 | ···················· | ···················· | ···················· | ···················· | ···················· | ···················· |
| W | 4177 | hCoV-19/England/ALDP-BBB50A/2020\|EPI_ISL_702010\|20 | ···················· | ···················· | ···················· | ···················· | ···················· | ···················· |
| W | 4178 | hCoV-19/England/ALDP-BBA107/2020\|EPI_ISL_702011\|20 | ···················· | ·········T·········· | ···················· | ···················· | ···················· | ···················· |
| W | 4179 | hCoV-19/England/ALDP-BB9E7A/2020\|EPI_ISL_702013\|20 | ········T··········· | ···················· | ···················· | ···················· | ···················· | ···················· |
| W | 4180 | hCoV-19/England/ALDP-BB37FD/2020\|EPI_ISL_702015\|20 | ···················· | ···················· | ···················· | ···················· | ···················· | ···················· |
| W | 4181 | hCoV-19/England/ALDP-BB360F/2020\|EPI_ISL_702016\|20 | ···················· | ···················· | ···················· | ···················· | ···················· | ···················· |
| W | 4182 | hCoV-19/England/ALDP-BB9FA4/2020\|EPI_ISL_702017\|20 | ···················· | ···················· | ···················· | ···················· | ···················· | ···················· |
| W | 4183 | hCoV-19/England/ALDP-BB5078/2020\|EPI_ISL_702018\|20 | ···················· | ·A·················· | ···················· | ···················· | ···················· | ···················· |
| W | 4184 | hCoV-19/England/ALDP-BBA76F/2020\|EPI_ISL_702019\|20 | ···················· | ···················· | ···················· | ···················· | ···················· | ···················· |
| W | 4185 | hCoV-19/England/ALDP-BB9E10/2020\|EPI_ISL_702020\|20 | ···················· | ···················· | ···················· | ···················· | ···················· | ···················· |
| W | 4186 | hCoV-19/England/ALDP-BBB018/2020\|EPI_ISL_702021\|20 | ···················· | ···················· | ···················· | ···················· | ···················· | ···················· |
| W | 4187 | hCoV-19/England/ALDP-BBA0DD/2020\|EPI_ISL_702022\|20 | ···················· | ···················· | ···················· | ···················· | ···················· | ···················· |
| W | 4188 | hCoV-19/England/CAMC-BD21EB/2020\|EPI_ISL_702025\|20 | ···················· | ···················· | ···················· | ···················· | ···················· | ···················· |
| W | 4189 | hCoV-19/England/CAMC-BD160B/2020\|EPI_ISL_702026\|20 | ···················· | ···················· | ···················· | ···················· | ···················· | ···················· |
| W | 4190 | hCoV-19/England/CAMC-BD22CA/2020\|EPI_ISL_702027\|20 | ···················· | ···················· | ···················· | ···················· | ···················· | ···················· |
| W | 4191 | hCoV-19/England/CAMC-BD13AA/2020\|EPI_ISL_702028\|20 | ···················· | ···················· | ···················· | ···················· | ···················· | ···················· |
| W | 4192 | hCoV-19/England/CAMC-BD4486/2020\|EPI_ISL_702029\|20 | ···················· | ···················· | ···················· | ···················· | ····T··············· | ···················· |
| W | 4193 | hCoV-19/England/CAMC-BD4608/2020\|EPI_ISL_702030\|20 | ···················· | ···················· | ···················· | ···················· | ······T············· | ···················· |
| W | 4194 | hCoV-19/England/CAMC-BD1EB8/2020\|EPI_ISL_702031\|20 | ···················· | ···················· | ···················· | ···················· | ···················· | ···················· |
| W | 4195 | hCoV-19/England/CAMC-BD2A0E/2020\|EPI_ISL_702032\|20 | ···················· | ···················· | ···················· | ···················· | ···················· | ···················· |
| W | 4196 | hCoV-19/England/CAMC-BD228E/2020\|EPI_ISL_702034\|20 | ···················· | ···················· | ···················· | ···················· | ···················· | ···················· |
| W | 4197 | hCoV-19/England/CAMC-BD485D/2020\|EPI_ISL_702038\|20 | ···················· | ···················· | ···················· | ···················· | ······T············· | ···················· |
| W | 4198 | hCoV-19/England/CAMC-BD2BDE/2020\|EPI_ISL_702040\|20 | ···················· | ···················· | ···················· | ···················· | ···················· | ···················· |
| W | 4199 | hCoV-19/England/CAMC-BD1586/2020\|EPI_ISL_702043\|20 | ···················· | ···················· | ···················· | ···················· | ···················· | ···················· |
| W | 4200 | hCoV-19/England/CAMC-BD12DA/2020\|EPI_ISL_702044\|20 | ···················· | ···················· | ···················· | ···················· | ···················· | ···················· |
| W | 4201 | hCoV-19/England/CAMC-BD1708/2020\|EPI_ISL_702045\|20 | ···················· | ···················· | ···················· | ···················· | ···················· | ···················· |
| W | 4202 | hCoV-19/England/CAMC-BD15C2/2020\|EPI_ISL_702051\|20 | ···················· | ···················· | ···················· | ···················· | ···················· | ···················· |
| W | 4203 | hCoV-19/England/CAMC-BD15B3/2020\|EPI_ISL_702052\|20 | ···················· | ···················· | ···················· | ···················· | ···················· | ···················· |
| W | 4204 | hCoV-19/England/CAMC-BD239A/2020\|EPI_ISL_702053\|20 | ···················· | ·A·················· | ···················· | ···················· | ···················· | ···A················ |
| W | 4205 | hCoV-19/England/CAMC-BD1E6D/2020\|EPI_ISL_702055\|20 | ···················· | ···················· | ···················· | ···················· | ······T············· | ···················· |
| W | 4206 | hCoV-19/England/CAMC-BD4626/2020\|EPI_ISL_702058\|20 | ···················· | ···················· | ···················· | ···················· | ···················· | ···················· |

| | | | | | | | |
|---|---|---|---|---|---|---|---|
| W | 4207 hCoV-19/England/CAMC-BD234F/2020\|EPI_ISL_702059\|20 | ···················· | ···················· | ···················· | ···················· | ···················· | ···················· |
| W | 4208 hCoV-19/England/CAMC-BD4644/2020\|EPI_ISL_702060\|20 | ···················· | ···················· | ···················· | ···················· | ···················· | ···················· |
| W | 4209 hCoV-19/England/CAMC-BD2AFF/2020\|EPI_ISL_702061\|20 | ···················· | ···················· | ···················· | ···················· | ···················· | ···················· |
| W | 4210 hCoV-19/England/CAMC-BD4617/2020\|EPI_ISL_702064\|20 | ···················· | ···················· | ···················· | ···················· | ···················· | ···················· |
| W | 4211 hCoV-19/England/CAMC-BD2B56/2020\|EPI_ISL_702067\|20 | ···················· | ···················· | ···················· | ···················· | ···················· | ···················· |
| W | 4212 hCoV-19/England/CAMC-BD17DB/2020\|EPI_ISL_702070\|20 | ···················· | ···················· | ···················· | ···················· | ···················· | ···················· |
| W | 4213 hCoV-19/England/CAMC-BD293E/2020\|EPI_ISL_702071\|20 | ···················· | ·A·················· | ···················· | ···················· | ···················· | ···················· |
| W | 4214 hCoV-19/England/CAMC-BD20C0/2020\|EPI_ISL_702076\|20 | ···················· | ···················· | ···················· | ···················· | ···················· | ···················· |
| W | 4215 hCoV-19/England/CAMC-BD2A2C/2020\|EPI_ISL_702077\|20 | ···················· | ···················· | ···················· | ···················· | ···················· | ···················· |
| W | 4216 hCoV-19/England/CAMC-BD1BC0/2020\|EPI_ISL_702079\|20 | ···················· | ···················· | ···················· | ···················· | ···················· | ···················· |
| W | 4217 hCoV-19/England/CAMC-BD1E5E/2020\|EPI_ISL_702080\|20 | ···················· | ···················· | ···················· | ···················· | ···················· | ···················· |
| W | 4218 hCoV-19/England/CAMC-BD4468/2020\|EPI_ISL_702084\|20 | ···················· | ···················· | ···················· | ···················· | ···················· | ···················· |
| W | 4219 hCoV-19/England/CAMC-BD2172/2020\|EPI_ISL_702087\|20 | ···················· | ···················· | ···················· | ···················· | ···················· | ···················· |
| W | 4220 hCoV-19/England/CAMC-BD23A9/2020\|EPI_ISL_702089\|20 | ···················· | ···················· | ···················· | ···················· | ···················· | ···················· |
| W | 4221 hCoV-19/England/CAMC-BD1F00/2020\|EPI_ISL_702091\|20 | ···················· | ···················· | ···················· | ···················· | ···················· | ···················· |
| W | 4222 hCoV-19/England/CAMC-BD1304/2020\|EPI_ISL_702099\|20 | ···················· | ···················· | ···················· | ···················· | ···················· | ···················· |
| W | 4223 hCoV-19/England/ALDP-BCA7E4/2020\|EPI_ISL_703311\|20 | ···················· | ···················· | ···················· | ···················· | ···················· | ···················· |
| W | 4224 hCoV-19/England/ALDP-BCA122/2020\|EPI_ISL_703312\|20 | ···················· | ···················· | ···················· | ···················· | ···················· | ···················· |
| W | 4225 hCoV-19/England/ALDP-BCAB06/2020\|EPI_ISL_703314\|20 | ···················· | ···················· | ···················· | ···················· | ···················· | ···A················ |
| W | 4226 hCoV-19/England/ALDP-BCA061/2020\|EPI_ISL_703317\|20 | ···················· | ···················· | ···················· | ···················· | ···················· | ···················· |
| W | 4227 hCoV-19/England/ALDP-BCAA36/2020\|EPI_ISL_703318\|20 | ···················· | ···················· | ···················· | ···················· | ···················· | ···················· |
| W | 4228 hCoV-19/England/ALDP-BCA26B/2020\|EPI_ISL_703341\|20 | ···················· | ···················· | ···················· | ···················· | ···················· | ···················· |
| W | 4229 hCoV-19/England/ALDP-BCABBB/2020\|EPI_ISL_703342\|20 | ···················· | ···················· | ···················· | ···················· | ···················· | ···················· |
| W | 4230 hCoV-19/England/QEUH-BC41BF/2020\|EPI_ISL_703345\|20 | ···················· | ···················· | ···················· | ···················· | ···················· | ···················· |
| W | 4231 hCoV-19/England/ALDP-BCA69C/2020\|EPI_ISL_703347\|20 | ···················· | ···················· | ···················· | ···················· | ···················· | ···················· |
| W | 4232 hCoV-19/England/QEUH-BC4234/2020\|EPI_ISL_703349\|20 | ···················· | ···················· | ···················· | ···················· | ······T············· | ···················· |
| W | 4233 hCoV-19/England/QEUH-BC429E/2020\|EPI_ISL_703350\|20 | ···················· | ···················· | ···················· | ···················· | ···················· | ···················· |
| W | 4234 hCoV-19/England/QEUH-BC4173/2020\|EPI_ISL_703352\|20 | ···················· | ···················· | ···················· | ···················· | ···················· | ···················· |
| W | 4235 hCoV-19/England/QEUH-BC41FB/2020\|EPI_ISL_703355\|20 | ···················· | ···················· | ···················· | ···················· | ···················· | ···················· |
| W | 4236 hCoV-19/England/ALDP-BCA386/2020\|EPI_ISL_703365\|20 | ···················· | ···················· | ···················· | ···················· | ···················· | ···················· |
| W | 4237 hCoV-19/England/ALDP-BCA517/2020\|EPI_ISL_703368\|20 | ···················· | ···················· | ···················· | ···················· | ···················· | ···················· |
| W | 4238 hCoV-19/England/QEUH-BC4094/2020\|EPI_ISL_703370\|20 | ···················· | ···················· | ···················· | ···················· | ······T············· | ···················· |
| W | 4239 hCoV-19/England/QEUH-BC4270/2020\|EPI_ISL_703371\|20 | ···················· | ···················· | ···················· | ···················· | ···················· | ···················· |
| W | 4240 hCoV-19/England/ALDP-BCA0BC/2020\|EPI_ISL_703374\|20 | ···················· | ···················· | ···················· | ···················· | ···················· | ···················· |
| W | 4241 hCoV-19/England/ALDP-BCA869/2020\|EPI_ISL_703376\|20 | ···················· | ···················· | ···················· | ···················· | ···················· | ···················· |
| W | 4242 hCoV-19/England/QEUH-BC47DB/2020\|EPI_ISL_703378\|20 | ···················· | ···················· | ···················· | ···················· | ···················· | ···················· |
| W | 4243 hCoV-19/England/QEUH-BC4629/2020\|EPI_ISL_703379\|20 | ···················· | ···················· | ···················· | ···················· | ···················· | ···················· |
| W | 4244 hCoV-19/England/QEUH-BC41CE/2020\|EPI_ISL_703381\|20 | ···················· | ···················· | ···················· | ···················· | ···················· | ···················· |
| W | 4245 hCoV-19/England/QEUH-BC44F2/2020\|EPI_ISL_703382\|20 | ···················· | ···················· | ···················· | ···················· | ···················· | ···················· |
| W | 4246 hCoV-19/England/QEUH-BC48BA/2020\|EPI_ISL_703384\|20 | ···················· | ···················· | ···················· | ···················· | ···················· | ···················· |
| W | 4247 hCoV-19/England/ALDP-BCA104/2020\|EPI_ISL_703386\|20 | ···················· | ···················· | ···················· | ···················· | ···················· | ···················· |
| W | 4248 hCoV-19/England/ALDP-BCA210/2020\|EPI_ISL_703388\|20 | ···················· | ···················· | ···················· | ···················· | ···················· | ···················· |
| W | 4249 hCoV-19/England/QEUH-BC4577/2020\|EPI_ISL_703389\|20 | ···················· | ···················· | ···················· | ···················· | ···················· | ···················· |
| W | 4250 hCoV-19/England/QEUH-BC4304/2020\|EPI_ISL_703391\|20 | ···················· | ···················· | ···················· | ···················· | ···················· | ···················· |
| W | 4251 hCoV-19/England/QEUH-BC3F6B/2020\|EPI_ISL_703393\|20 | ···················· | ·A·················· | ···················· | ···················· | ···················· | ···A················ |
| W | 4252 hCoV-19/England/ALDP-BCA447/2020\|EPI_ISL_703394\|20 | ···················· | ···················· | ···················· | ···················· | ···················· | ···················· |
| W | 4253 hCoV-19/England/ALDP-BCAB8E/2020\|EPI_ISL_703396\|20 | ···················· | ···················· | ···················· | ···················· | ···················· | ···················· |
| W | 4254 hCoV-19/England/ALDP-BCA025/2020\|EPI_ISL_703397\|20 | ···················· | ···················· | ···················· | ···················· | ···················· | ···················· |
| W | 4255 hCoV-19/England/QEUH-BC4191/2020\|EPI_ISL_703399\|20 | ···················· | ···················· | ···················· | ···················· | ···················· | ···················· |
| W | 4256 hCoV-19/England/QEUH-BCDA33/2020\|EPI_ISL_704642\|20 | ···················· | ···················· | ···················· | ···················· | ···················· | ···················· |
| W | 4257 hCoV-19/England/QEUH-BCBA71/2020\|EPI_ISL_704644\|20 | ···················· | ···················· | ···················· | ···················· | ···················· | ···················· |
| W | 4258 hCoV-19/England/QEUH-BCCABC/2020\|EPI_ISL_704645\|20 | ···················· | ···················· | ···················· | ···················· | ···················· | ···················· |
| W | 4259 hCoV-19/England/QEUH-BCC1E4/2020\|EPI_ISL_704647\|20 | ···················· | ···················· | ···················· | ···················· | ···················· | ···················· |
| W | 4260 hCoV-19/England/QEUH-BCBC4E/2020\|EPI_ISL_704649\|20 | ···················· | ···················· | ···················· | ···················· | ······T············· | ···················· |
| W | 4261 hCoV-19/England/QEUH-BCCAAD/2020\|EPI_ISL_704670\|20 | ···················· | ···················· | ···················· | ···················· | ···················· | ···················· |
| W | 4262 hCoV-19/England/QEUH-BCE2B2/2020\|EPI_ISL_704672\|20 | ···················· | ···················· | ···················· | ···················· | ···················· | ···················· |
| W | 4263 hCoV-19/England/QEUH-BCBA44/2020\|EPI_ISL_704673\|20 | ···················· | ···················· | ···················· | ···················· | ···················· | ···················· |
| W | 4264 hCoV-19/England/QEUH-BCD82A/2020\|EPI_ISL_704675\|20 | ···················· | ···················· | ···················· | ···················· | ···················· | ···················· |
| W | 4265 hCoV-19/England/QEUH-BCD4BD/2020\|EPI_ISL_704678\|20 | ···················· | ···················· | ···················· | ···················· | ···················· | ···················· |
| W | 4266 hCoV-19/England/QEUH-BCC560/2020\|EPI_ISL_704680\|20 | ···················· | ···················· | ···················· | ···················· | ···················· | ···················· |
| W | 4267 hCoV-19/England/QEUH-BCD7F0/2020\|EPI_ISL_704681\|20 | ···················· | ···················· | ···················· | ···················· | ···················· | ···················· |
| W | 4268 hCoV-19/England/QEUH-BCCF71/2020\|EPI_ISL_704684\|20 | ···················· | ···················· | ···················· | ···················· | ···················· | ···················· |
| W | 4269 hCoV-19/England/QEUH-BCC18A/2020\|EPI_ISL_704686\|20 | ···················· | ···················· | ···················· | ···················· | ···················· | ···················· |
| W | 4270 hCoV-19/England/CAMC-BC00F2/2020\|EPI_ISL_704688\|20 | ···················· | ···················· | ···················· | ···················· | ···················· | ···················· |
| W | 4271 hCoV-19/England/CAMC-BC0362/2020\|EPI_ISL_704689\|20 | ···················· | ·A·················· | ···················· | ···················· | ···················· | ···················· |
| W | 4272 hCoV-19/England/CAMC-BC1A3F/2020\|EPI_ISL_704691\|20 | ···················· | ···················· | ···················· | ···················· | ···················· | ···················· |
| W | 4273 hCoV-19/England/CAMC-BBFF53/2020\|EPI_ISL_704694\|20 | ···················· | ···················· | ···················· | ···················· | ······T············· | ···················· |
| W | 4274 hCoV-19/England/CAMC-BC1C39/2020\|EPI_ISL_704696\|20 | ···················· | ···················· | ···················· | ···················· | ···················· | ···················· |
| W | 4275 hCoV-19/England/CAMC-BC014A/2020\|EPI_ISL_704697\|20 | ········T··········· | ···················· | ···················· | ···················· | ···················· | ···················· |
| W | 4276 hCoV-19/England/LOND-12646C7/2020\|EPI_ISL_705970\|2 | ···················· | ···················· | ···················· | ···················· | ···················· | ···················· |
| W | 4277 hCoV-19/England/LOND-126475B/2020\|EPI_ISL_705971\|2 | ···················· | ···················· | ···················· | ···················· | ···················· | ···················· |
| W | 4278 hCoV-19/England/MILK-A91680/2020\|EPI_ISL_705974\|20 | ···················· | ···················· | ···················· | ···················· | ···················· | ···················· |
| W | 4279 hCoV-19/England/MILK-A916AE/2020\|EPI_ISL_705975\|20 | ···················· | ···················· | ···················· | ···················· | ···················· | ···················· |
| W | 4280 hCoV-19/England/MILK-A916F9/2020\|EPI_ISL_705976\|20 | ···················· | ···················· | ···················· | ···················· | ···················· | ···················· |
| W | 4281 hCoV-19/England/MILK-A91705/2020\|EPI_ISL_705977\|20 | ···················· | ···················· | ···················· | ···················· | ···················· | ···················· |
| W | 4282 hCoV-19/England/MILK-A91714/2020\|EPI_ISL_705978\|20 | ···················· | ···················· | ···················· | ···················· | ···················· | ···················· |
| W | 4283 hCoV-19/England/MILK-A91723/2020\|EPI_ISL_705979\|20 | ···················· | ···················· | ···················· | ···················· | ···················· | ···················· |
| W | 4284 hCoV-19/USA/MI-MDHHS-SC22501/2020\|EPI_ISL_708204\|2 | ···················· | ···················· | ···················· | ···················· | ···················· | ···················· |
| W | 4285 hCoV-19/USA/MI-MDHHS-SC22502/2020\|EPI_ISL_708205\|2 | ···················· | ···················· | ···················· | ···················· | ···················· | ···················· |
| W | 4286 hCoV-19/USA/MI-MDHHS-SC22504/2020\|EPI_ISL_708207\|2 | ···················· | ···················· | ···················· | ···················· | ···················· | ···················· |

| | | | | | | | | |
|---|---|---|---|---|---|---|---|---|
| W | 4287 | hCoV-19/USA/MI-MDHHS-SC22505/2020\|EPI_ISL_708208\|2 | ···················· | ···················· | ····················· | ···················· | ···················· | ···················· |
| W | 4288 | hCoV-19/USA/MI-MDHHS-SC22506/2020\|EPI_ISL_708209\|2 | ···················· | ···················· | ····················· | ···················· | ···················· | ···················· |
| W | 4289 | hCoV-19/USA/MI-MDHHS-SC22507/2020\|EPI_ISL_708210\|2 | ···················· | ···················· | ····················· | ···················· | ···················· | ···················· |
| W | 4290 | hCoV-19/USA/MI-MDHHS-SC22508/2020\|EPI_ISL_708211\|2 | ···················· | ···················· | ····················· | ···················· | ···················· | ···················· |
| W | 4291 | hCoV-19/USA/MI-MDHHS-SC22509/2020\|EPI_ISL_708212\|2 | ···················· | ···················· | ····················· | ···················· | ···················· | ···················· |
| W | 4292 | hCoV-19/USA/MI-MDHHS-SC22510/2020\|EPI_ISL_708213\|2 | ···················· | ···················· | ····················· | ···················· | ···················· | ···················· |
| W | 4293 | hCoV-19/USA/MI-MDHHS-SC22511/2020\|EPI_ISL_708214\|2 | ···················· | ···················· | ····················· | ···················· | ···················· | ···················· |
| W | 4294 | hCoV-19/USA/MI-MDHHS-SC22512/2020\|EPI_ISL_708215\|2 | ···················· | ···················· | ····················· | ···················· | ···················· | ···················· |
| W | 4295 | hCoV-19/USA/MI-MDHHS-SC22513/2020\|EPI_ISL_708216\|2 | ···················· | ···················· | ····················· | ···················· | ···················· | ···················· |
| W | 4296 | hCoV-19/USA/MI-MDHHS-SC22514/2020\|EPI_ISL_708217\|2 | ···················· | ···················· | ····················· | ···················· | ···················· | ···················· |
| W | 4297 | hCoV-19/USA/MI-MDHHS-SC22515/2020\|EPI_ISL_708218\|2 | ···················· | ···················· | ····················· | ···················· | ···················· | ···················· |
| W | 4298 | hCoV-19/USA/MI-MDHHS-SC22516/2020\|EPI_ISL_708219\|2 | ···················· | ···················· | ····················· | ···················· | ···················· | ···················· |
| W | 4299 | hCoV-19/USA/MI-MDHHS-SC22517/2020\|EPI_ISL_708220\|2 | ···················· | ···················· | ····················· | ···················· | ···················· | ···················· |
| W | 4300 | hCoV-19/USA/MI-MDHHS-SC22518/2020\|EPI_ISL_708221\|2 | ···················· | ···················· | ····················· | ···················· | ···················· | ···················· |
| W | 4301 | hCoV-19/USA/MI-MDHHS-SC22519/2020\|EPI_ISL_708222\|2 | ···················· | ···················· | ····················· | ···················· | ···················· | ···················· |
| W | 4302 | hCoV-19/USA/MI-MDHHS-SC22520/2020\|EPI_ISL_708223\|2 | ···················· | ···················· | ····················· | ···················· | ···················· | ···················· |
| W | 4303 | hCoV-19/USA/MI-MDHHS-SC22521/2020\|EPI_ISL_708224\|2 | ···················· | ···················· | ····················· | ···················· | ···················· | ···················· |
| W | 4304 | hCoV-19/USA/MI-MDHHS-SC22522/2020\|EPI_ISL_708225\|2 | ···················· | ···················· | ····················· | ···················· | ···················· | ···················· |
| W | 4305 | hCoV-19/USA/MI-MDHHS-SC22523/2020\|EPI_ISL_708226\|2 | ···················· | ···················· | ····················· | ···················· | ···················· | ···················· |
| W | 4306 | hCoV-19/USA/MI-MDHHS-SC22524/2020\|EPI_ISL_708227\|2 | ···················· | ···················· | ····················· | ···················· | ···················· | ···················· |
| W | 4307 | hCoV-19/USA/MI-MDHHS-SC22525/2020\|EPI_ISL_708228\|2 | ···················· | ···················· | ····················· | ···················· | ···················· | ···················· |
| W | 4308 | hCoV-19/USA/MI-MDHHS-SC22526/2020\|EPI_ISL_708229\|2 | ···················· | ···················· | ····················· | ···················· | ···················· | ···················· |
| W | 4309 | hCoV-19/USA/MI-MDHHS-SC22417/2020\|EPI_ISL_708250\|2 | ···················· | ···················· | ····················· | ···················· | ···················· | ···················· |
| W | 4310 | hCoV-19/USA/MI-MDHHS-SC22418/2020\|EPI_ISL_708251\|2 | ···················· | ···················· | ····················· | ···················· | ···················· | ···················· |
| W | 4311 | hCoV-19/USA/MI-MDHHS-SC22419/2020\|EPI_ISL_708252\|2 | ···················· | ···················· | ····················· | ···················· | ···················· | ···················· |
| W | 4312 | hCoV-19/USA/MI-MDHHS-SC22420/2020\|EPI_ISL_708253\|2 | ···················· | ···················· | ····················· | ···················· | ···················· | ···················· |
| W | 4313 | hCoV-19/England/QEUH-C15F78/2020\|EPI_ISL_709500\|20 | ···················· | ···················· | ····················· | ···················· | ·····T·············· | ···················· |
| W | 4314 | hCoV-19/England/ALDP-C060A4/2020\|EPI_ISL_709501\|20 | ···················· | ···················· | ····················· | ···················· | ···················· | ···················· |
| W | 4315 | hCoV-19/England/QEUH-C14E8B/2020\|EPI_ISL_709502\|20 | ···················· | ···················· | ····················· | ···················· | ···················· | ···················· |
| W | 4316 | hCoV-19/England/QEUH-C14C27/2020\|EPI_ISL_709503\|20 | ···················· | ···················· | ····················· | ···················· | ···················· | ···················· |
| W | 4317 | hCoV-19/England/ALDP-C064A8/2020\|EPI_ISL_709504\|20 | ···················· | ···················· | ····················· | ···················· | ···················· | ···················· |
| W | 4318 | hCoV-19/England/ALDP-C06217/2020\|EPI_ISL_709507\|20 | ···················· | ···················· | ····················· | ···················· | ···················· | ···················· |
| W | 4319 | hCoV-19/England/ALDP-C0648A/2020\|EPI_ISL_709508\|20 | ···················· | ···················· | ····················· | ···················· | ···················· | ···················· |
| W | 4320 | hCoV-19/England/QEUH-C14BC0/2020\|EPI_ISL_709509\|20 | ···················· | ···················· | ····················· | ···················· | ···················· | ···················· |
| W | 4321 | hCoV-19/England/QEUH-C1597A/2020\|EPI_ISL_709510\|20 | ···················· | ···················· | ····················· | ···················· | ···················· | ···················· |
| W | 4322 | hCoV-19/England/ALDP-C06824/2020\|EPI_ISL_709512\|20 | ···················· | ···················· | ····················· | ···················· | ···················· | ···················· |
| W | 4323 | hCoV-19/England/ALDP-C061DE/2020\|EPI_ISL_709514\|20 | ···················· | ···················· | ····················· | ···················· | ···················· | ···················· |
| W | 4324 | hCoV-19/England/ALDP-C064D5/2020\|EPI_ISL_709516\|20 | ···················· | ···················· | ····················· | ···················· | ···················· | ···················· |
| W | 4325 | hCoV-19/England/QEUH-C145E0/2020\|EPI_ISL_709517\|20 | ···················· | ···················· | ····················· | ···················· | ···················· | ···················· |
| W | 4326 | hCoV-19/England/QEUH-C15F5A/2020\|EPI_ISL_709518\|20 | ···················· | ···················· | ····················· | ···················· | ···················· | ···················· |
| W | 4327 | hCoV-19/England/ALDP-C06402/2020\|EPI_ISL_709519\|20 | ···················· | ···················· | ····················· | ···················· | ···················· | ···················· |
| W | 4328 | hCoV-19/England/ALDP-C064B7/2020\|EPI_ISL_709521\|20 | ···················· | ···················· | ····················· | ···················· | ···················· | ···················· |
| W | 4329 | hCoV-19/England/QEUH-C1452C/2020\|EPI_ISL_709522\|20 | ···················· | ···················· | ····················· | ···················· | ···················· | ···················· |
| W | 4330 | hCoV-19/England/ALDP-C0653C/2020\|EPI_ISL_709523\|20 | ···················· | ···················· | ····················· | ···················· | ···················· | ···················· |
| W | 4331 | hCoV-19/England/QEUH-C14683/2020\|EPI_ISL_709524\|20 | ···················· | ···················· | ····················· | ···················· | ···················· | ···················· |
| W | 4332 | hCoV-19/England/QEUH-C14B93/2020\|EPI_ISL_709527\|20 | ···················· | ···················· | ····················· | ···················· | ···················· | ···················· |
| W | 4333 | hCoV-19/England/QEUH-C14BB1/2020\|EPI_ISL_709530\|20 | ···················· | ···················· | ····················· | ···················· | ···················· | ···················· |
| W | 4334 | hCoV-19/England/QEUH-C14629/2020\|EPI_ISL_709533\|20 | ···················· | ···················· | ····················· | ···················· | ···················· | ···················· |
| W | 4335 | hCoV-19/England/QEUH-C14B84/2020\|EPI_ISL_709537\|20 | ···················· | ···················· | ····················· | ···················· | ···················· | ···················· |
| W | 4336 | hCoV-19/England/QEUH-C1640F/2020\|EPI_ISL_709539\|20 | ···················· | ···················· | ····················· | ···················· | ···················· | ···················· |
| W | 4337 | hCoV-19/England/QEUH-C1453B/2020\|EPI_ISL_709541\|20 | ···················· | ···················· | ····················· | ···················· | ···················· | ···················· |
| W | 4338 | hCoV-19/England/ALDP-C066C0/2020\|EPI_ISL_709543\|20 | ···················· | ···················· | ····················· | ···················· | ···················· | ···················· |
| W | 4339 | hCoV-19/England/ALDP-C06068/2020\|EPI_ISL_709544\|20 | ···················· | ···················· | ····················· | ···················· | ···················· | ···················· |
| W | 4340 | hCoV-19/England/ALDP-C068D9/2020\|EPI_ISL_709545\|20 | ···················· | ···················· | ····················· | ···················· | ···················· | ···················· |
| W | 4341 | hCoV-19/England/ALDP-C06DCB/2020\|EPI_ISL_709546\|20 | ···················· | ···················· | ····················· | ···················· | ···················· | ···················· |
| W | 4342 | hCoV-19/England/QEUH-C16074/2020\|EPI_ISL_709549\|20 | ···················· | ···················· | ····················· | ···················· | ···················· | ···················· |
| W | 4343 | hCoV-19/England/ALDP-C05F99/2020\|EPI_ISL_709550\|20 | ···················· | ···················· | ····················· | ···················· | ···················· | ···················· |
| W | 4344 | hCoV-19/England/ALDP-C060C2/2020\|EPI_ISL_709552\|20 | ···················· | ···················· | ····················· | ···················· | ···················· | ···················· |
| W | 4345 | hCoV-19/England/ALDP-C0660C/2020\|EPI_ISL_709554\|20 | ···················· | ···················· | ····················· | ···················· | ···················· | ···················· |
| W | 4346 | hCoV-19/England/QEUH-C15A1D/2020\|EPI_ISL_709555\|20 | ···················· | ···················· | ····················· | ···················· | ···················· | ···················· |
| W | 4347 | hCoV-19/England/QEUH-C14A1E/2020\|EPI_ISL_709557\|20 | ···················· | ···················· | ····················· | ···················· | ···················· | ···················· |
| W | 4348 | hCoV-19/England/QEUH-C1460B/2020\|EPI_ISL_709558\|20 | ···················· | ···················· | ····················· | ···················· | ···················· | ···················· |
| W | 4349 | hCoV-19/England/ALDP-C06095/2020\|EPI_ISL_709559\|20 | ···················· | ···················· | ····················· | ···················· | ···················· | ···················· |
| W | 4350 | hCoV-19/England/MILK-BDC6B5/2020\|EPI_ISL_709582\|20 | ···················· | ···················· | ····················· | ···················· | ···················· | ···················· |
| W | 4351 | hCoV-19/England/MILK-BDC196/2020\|EPI_ISL_709583\|20 | ···················· | ···················· | ····················· | ···················· | ···················· | ···················· |
| W | 4352 | hCoV-19/England/MILK-BDCB2F/2020\|EPI_ISL_709584\|20 | ···················· | ···················· | ····················· | ···················· | ···················· | ···················· |
| W | 4353 | hCoV-19/USA/AZ-TG366750/2020\|EPI_ISL_694503\|2020-0 | ···················· | ···················· | ····················· | ···················· | ···················· | ···················· |
| W | 4354 | hCoV-19/USA/AZ-TG366762/2020\|EPI_ISL_694509\|2020-0 | ···················· | ···················· | ····················· | ···················· | ···················· | ···················· |
| W | 4355 | hCoV-19/USA/AZ-TG371307/2020\|EPI_ISL_694560\|2020-0 | ···················· | ···················· | ····················· | ···················· | ···················· | ···················· |
| W | 4356 | hCoV-19/USA/AZ-TG371309/2020\|EPI_ISL_694562\|2020-0 | ···················· | ···················· | ····················· | ···················· | ···················· | ···················· |
| W | 4357 | hCoV-19/USA/AZ-TG371315/2020\|EPI_ISL_694567\|2020-0 | ···················· | ·············A······ | ····················· | ···················· | ···················· | ···················· |
| W | 4358 | hCoV-19/USA/AZ-TG371318/2020\|EPI_ISL_694569\|2020-0 | ···················· | ···················· | ····················· | ···················· | ···················· | ···················· |
| W | 4359 | hCoV-19/USA/AZ-TG371322/2020\|EPI_ISL_694572\|2020-0 | ···················· | ···················· | ····················· | ···················· | ···················· | ···················· |
| W | 4360 | hCoV-19/USA/AZ-TG371324/2020\|EPI_ISL_694574\|2020-0 | ···················· | ···················· | ····················· | ···················· | ···················· | ···················· |
| W | 4361 | hCoV-19/USA/AZ-TG371332/2020\|EPI_ISL_694576\|2020-0 | ···········T········ | ···················· | ····················· | ···················· | ···················· | ···················· |
| W | 4362 | hCoV-19/USA/AZ-TG371333/2020\|EPI_ISL_694577\|2020-0 | ···········T········ | ···················· | ····················· | ···················· | ···················· | ···················· |
| W | 4363 | hCoV-19/USA/AZ-TG371334/2020\|EPI_ISL_694578\|2020-0 | ···········T········ | ···················· | ····················· | ···················· | ···················· | ···················· |
| W | 4364 | hCoV-19/USA/AZ-TG371335/2020\|EPI_ISL_694579\|2020-0 | ···········T········ | ···················· | ····················· | ···················· | ···················· | ···················· |
| W | 4365 | hCoV-19/USA/AZ-TG371336/2020\|EPI_ISL_694580\|2020-0 | ···········T········ | ···················· | ····················· | ···················· | ···················· | ···················· |
| W | 4366 | hCoV-19/USA/AZ-TG371337/2020\|EPI_ISL_694581\|2020-0 | ···········T········ | ···················· | ····················· | ···················· | ···················· | ···················· |

W 4367 hCoV-19/USA/AZ-TG371338/2020|EPI_ISL_694582|2020-0 ···········T········ ···················· ····················· ···················· ···················· ····················
W 4368 hCoV-19/USA/AZ-TG371341/2020|EPI_ISL_694585|2020-0 ···········T········ ···················· ····················· ···················· ···················· ····················
W 4369 hCoV-19/USA/AZ-TG371345/2020|EPI_ISL_694588|2020-0 ···················· ···················· ····················· ···················· ···················· ····················
W 4370 hCoV-19/USA/AZ-TG371347/2020|EPI_ISL_694589|2020-0 ···················· ···················· ····················· ···················· ···················· ····················
W 4371 hCoV-19/USA/AZ-TG371349/2020|EPI_ISL_694591|2020-0 ···················· ···················· ····················· ···················· ···················· ····················
W 4372 hCoV-19/USA/AZ-TG371350/2020|EPI_ISL_694592|2020-0 ···················· ···················· ····················· ···················· ···················· ····················
W 4373 hCoV-19/USA/AZ-TG371353/2020|EPI_ISL_694595|2020-0 ···················· ···················· ····················· ···················· ···················· ····················
W 4374 hCoV-19/USA/AZ-TG371355/2020|EPI_ISL_694596|2020-0 ···················· ···················· ····················· ···················· ···················· ····················
W 4375 hCoV-19/USA/AZ-TG640868/2020|EPI_ISL_695802|2020-0 ···················· ···················· ····················· ···················· ···················· ····················
W 4376 hCoV-19/USA/AZ-TG640871/2020|EPI_ISL_695803|2020-0 ···················· ···················· ····················· ···················· ···················· ····················
W 4377 hCoV-19/USA/AZ-TG640872/2020|EPI_ISL_695804|2020-0 ···················· ···················· ····················· ···················· ···················· ····················
W 4378 hCoV-19/USA/AZ-TG640908/2020|EPI_ISL_695812|2020-0 ···················· ···················· ····················· ···················· ···················· ····················
W 4379 hCoV-19/USA/AZ-TG640916/2020|EPI_ISL_695815|2020-0 ···················· ···················· ····················· ···················· ···················· ····················
W 4380 hCoV-19/USA/AZ-TG640922/2020|EPI_ISL_695817|2020-0 ···················· ···················· ····················· ···················· ···················· ····················
W 4381 hCoV-19/USA/AZ-TG640927/2020|EPI_ISL_695819|2020-0 ···················· ···················· ····················· ···················· ···················· ····················
W 4382 hCoV-19/USA/AZ-TG640944/2020|EPI_ISL_695822|2020-0 ···················· ···················· ····················· ···················· ···················· ····················
W 4383 hCoV-19/USA/AZ-TG640950/2020|EPI_ISL_695823|2020-0 ···················· ···················· ····················· ···················· ···················· ····················
W 4384 hCoV-19/USA/AZ-TG640956/2020|EPI_ISL_695824|2020-0 ···················· ···················· ····················· ···················· ···················· ····················
W 4385 hCoV-19/USA/AZ-TG640957/2020|EPI_ISL_695825|2020-0 ···················· ···················· ····················· ···················· ···················· ····················
W 4386 hCoV-19/USA/AZ-TG640962/2020|EPI_ISL_695826|2020-0 ···················· ···················· ····················· ···················· ···················· ····················
W 4387 hCoV-19/USA/AZ-TG640965/2020|EPI_ISL_695828|2020-0 ···················· ···················· ····················· ···················· ···················· ····················
W 4388 hCoV-19/USA/AZ-TG640967/2020|EPI_ISL_695829|2020-0 ···················· ···················· ····················· ···················· ···················· ····················
W 4389 hCoV-19/USA/AZ-TG645812/2020|EPI_ISL_695830|2020-0 ···················· ·A·················· ····················· ···················· ···················· ····················
W 4390 hCoV-19/USA/AZ-TG648694/2020|EPI_ISL_695896|2020-0 ···················· ···················· ····················· ···················· ···················· ····················
W 4391 hCoV-19/Canada/ON-UHTC_0398/2020|EPI_ISL_698079|20 ···················· ···················· ····················· ···················· ···················· ····················
W 4392 hCoV-19/Canada/ON-UHTC_0399/2020|EPI_ISL_698080|20 ···················· ···················· ····················· ···················· ···················· ····················
W 4393 hCoV-19/Canada/ON-UHTC_0395/2020|EPI_ISL_698081|20 ···················· ·A·················· ····················· ···················· ···················· ···A················
W 4394 hCoV-19/Canada/ON-UHTC_0406/2020|EPI_ISL_698083|20 ···················· ·A·················· ····················· ···················· ···················· ···A················
W 4395 hCoV-19/Canada/ON-UHTC_0405/2020|EPI_ISL_698084|20 ···················· ·A·················· ····················· ···················· ···················· ···A················
W 4396 hCoV-19/Canada/ON-UHTC_0407/2020|EPI_ISL_698085|20 ···················· ···················· ····················· ···················· ···················· ····················
W 4397 hCoV-19/Canada/ON-UHTC_0393/2020|EPI_ISL_698086|20 ···················· ···················· ····················· ···················· ···················· ····················
W 4398 hCoV-19/Canada/ON-UHTC_0401/2020|EPI_ISL_698089|20 ···················· ···················· ····················· ···················· ···················· ····················
W 4399 hCoV-19/Canada/ON-UHTC_0397/2020|EPI_ISL_698090|20 ···················· ···················· ····················· ···················· ···················· ····················
W 4400 hCoV-19/Canada/ON-UHTC_0388/2020|EPI_ISL_698091|20 ···················· ···················· ····················· ···················· ···················· ····················
W 4401 hCoV-19/Canada/ON-UHTC_0396/2020|EPI_ISL_698092|20 ···················· ···················· ····················· ···················· ···················· ····················
W 4402 hCoV-19/Canada/ON-UHTC_0409/2020|EPI_ISL_698093|20 ···················· ·A·················· ····················· ···················· ···················· ···A················
W 4403 hCoV-19/Canada/ON-UHTC_0382/2020|EPI_ISL_698094|20 ···················· ·A·················· ····················· ···················· ···················· ···A················
W 4404 hCoV-19/Canada/ON-UHTC_0389/2020|EPI_ISL_698095|20 ···················· ·A·················· ····················· ···················· ···················· ···A················
W 4405 hCoV-19/Canada/ON-UHTC_0391/2020|EPI_ISL_698096|20 ···················· ·A·················· ····················· ···················· ···················· ···A················
W 4406 hCoV-19/Canada/ON-UHTC_0400/2020|EPI_ISL_698097|20 ···················· ···················· ····················· ···················· ···················· ····················
W 4407 hCoV-19/Canada/ON-UHTC_0394/2020|EPI_ISL_698098|20 ···················· ···················· ····················· ···················· ···················· ····················
W 4408 hCoV-19/Canada/ON-UHTC_0384/2020|EPI_ISL_698099|20 ···················· ·A·················· ····················· ···················· ···················· ···A················
W 4409 hCoV-19/England/CAMC-BD2163/2020|EPI_ISL_702101|20 ···················· ···················· ····················· ···················· ···················· ····················
W 4410 hCoV-19/England/CAMC-BD1B1B/2020|EPI_ISL_702104|20 ···················· ···················· ····················· ···················· ···················· ····················
W 4411 hCoV-19/England/CAMC-BD135F/2020|EPI_ISL_702106|20 ···················· ···················· ····················· ···················· ···················· ····················
W 4412 hCoV-19/England/CAMC-BD20DF/2020|EPI_ISL_702107|20 ···················· ···················· ····················· ···················· ···················· ····················
W 4413 hCoV-19/England/CAMC-BD2127/2020|EPI_ISL_702108|20 ···················· ···················· ····················· ···················· ···················· ····················
W 4414 hCoV-19/England/CAMC-BD48D5/2020|EPI_ISL_702113|20 ···················· ···················· ····················· ···················· ···················· ····················
W 4415 hCoV-19/England/CAMC-BD1EF4/2020|EPI_ISL_702114|20 ···················· ···················· ····················· ···················· ···················· ····················
W 4416 hCoV-19/England/CAMC-BD1410/2020|EPI_ISL_702115|20 ···················· ···················· ····················· ···················· ···················· ····················
W 4417 hCoV-19/England/CAMC-BD469F/2020|EPI_ISL_702118|20 ···················· ···················· ····················· ···················· ···················· ····················
W 4418 hCoV-19/England/CAMC-BD2057/2020|EPI_ISL_702120|20 ···················· ···················· ····················· ···················· ···················· ····················
W 4419 hCoV-19/England/CAMC-BD1F79/2020|EPI_ISL_702121|20 ···················· ···················· ····················· ···················· ···················· ····················
W 4420 hCoV-19/England/CAMC-BD1243/2020|EPI_ISL_702122|20 ···················· ···················· ····················· ···················· ···················· ····················
W 4421 hCoV-19/England/CAMC-BD1577/2020|EPI_ISL_702123|20 ···················· ···················· ····················· ···················· ···················· ····················
W 4422 hCoV-19/England/CAMC-BD22F7/2020|EPI_ISL_702124|20 ···················· ···················· ····················· ···················· ···················· ····················
W 4423 hCoV-19/England/CAMC-BD4680/2020|EPI_ISL_702125|20 ···················· ···················· ····················· ···················· ···················· ····················
W 4424 hCoV-19/England/CAMC-BD2B38/2020|EPI_ISL_702127|20 ···················· ···················· ····················· ···················· ···················· ····················
W 4425 hCoV-19/England/CAMC-BD4398/2020|EPI_ISL_702128|20 ···················· ···················· ····················· ···················· ···················· ····················
W 4426 hCoV-19/England/CAMC-BD46AE/2020|EPI_ISL_702129|20 ···················· ···················· ····················· ···················· ···················· ····················
W 4427 hCoV-19/England/CAMC-BD45FC/2020|EPI_ISL_702131|20 ···················· ···················· ····················· ···················· ···················· ····················
W 4428 hCoV-19/England/CAMC-BD1A3C/2020|EPI_ISL_702133|20 ···················· ···················· ····················· ···················· ···················· ····················
W 4429 hCoV-19/England/CAMC-BD43F2/2020|EPI_ISL_702135|20 ···················· ···················· ····················· ···················· ···················· ····················
W 4430 hCoV-19/England/CAMC-BD1A4B/2020|EPI_ISL_702136|20 ···················· ·A·················· ····················· ···················· ···················· ···A················
W 4431 hCoV-19/England/CAMC-BD12CB/2020|EPI_ISL_702139|20 ···················· ···················· ····················· ···················· ···················· ····················
W 4432 hCoV-19/England/CAMC-BD16CF/2020|EPI_ISL_702140|20 ···················· ···················· ····················· ···················· ···················· ····················
W 4433 hCoV-19/England/CAMC-BD1A1E/2020|EPI_ISL_702143|20 ···················· ···················· ····················· ···················· ···················· ····················
W 4434 hCoV-19/England/CAMC-BD46DB/2020|EPI_ISL_702150|20 ···················· ···················· ····················· ···················· ···················· ····················
W 4435 hCoV-19/England/CAMC-BD1A96/2020|EPI_ISL_702152|20 ···················· ·A·················· ····················· ···················· ···················· ···A················
W 4436 hCoV-19/England/CAMC-BD4635/2020|EPI_ISL_702153|20 ···················· ···················· ····················· ···················· ···················· ····················
W 4437 hCoV-19/England/CAMC-BD1832/2020|EPI_ISL_702157|20 ···················· ···················· ····················· ···················· ···················· ····················
W 4438 hCoV-19/England/CAMC-BD2066/2020|EPI_ISL_702158|20 ···················· ···················· ····················· ···················· ···················· ····················
W 4439 hCoV-19/England/CAMC-BD21AF/2020|EPI_ISL_702159|20 ···················· ···················· ····················· ···················· ···················· ····················
W 4440 hCoV-19/England/CAMC-BD1A87/2020|EPI_ISL_702160|20 ···················· ···················· ····················· ···················· ···················· ····················
W 4441 hCoV-19/England/CAMC-BD1C90/2020|EPI_ISL_702161|20 ···················· ···················· ····················· ···················· ···················· ····················
W 4442 hCoV-19/England/CAMC-BD21BE/2020|EPI_ISL_702163|20 ···················· ···················· ····················· ···················· ···················· ····················
W 4443 hCoV-19/England/CAMC-BD1656/2020|EPI_ISL_702164|20 ···················· ···················· ····················· ···················· ···················· ····················
W 4444 hCoV-19/England/CAMC-BD15A4/2020|EPI_ISL_702166|20 ···················· ···················· ····················· ···················· ···················· ····················
W 4445 hCoV-19/England/QEUH-BC3F01/2020|EPI_ISL_703401|20 ···················· ···················· ····················· ···················· ···················· ····················
W 4446 hCoV-19/England/ALDP-BCA1B9/2020|EPI_ISL_703402|20 ···················· ···················· ····················· ···················· ···················· ····················

W 4447 hCoV-19/England/ALDP-BCA614/2020|EPI_ISL_703404|20 ···················· ···················· ····················· ···················· ···················· ····················
W 4448 hCoV-19/England/ALDP-BCA25C/2020|EPI_ISL_703405|20 ···················· ···················· ····················· ···················· ······T············· ····················
W 4449 hCoV-19/England/QEUH-BC40C1/2020|EPI_ISL_703407|20 ···················· ···················· ····················· ···················· ···················· ····················
W 4450 hCoV-19/England/QEUH-BC4155/2020|EPI_ISL_703408|20 ···················· ···················· ····················· ···················· ···················· ····················
W 4451 hCoV-19/England/QEUH-BC3EC8/2020|EPI_ISL_703410|20 ···················· ···················· ····················· ···················· ···················· ····················
W 4452 hCoV-19/England/QEUH-BC42DA/2020|EPI_ISL_703411|20 ···················· ···················· ····················· ···················· ···················· ····················
W 4453 hCoV-19/England/ALDP-BCA16E/2020|EPI_ISL_703413|20 ···················· ···················· ····················· ···················· ···················· ····················
W 4454 hCoV-19/England/QEUH-BC4735/2020|EPI_ISL_703415|20 ···················· ···················· ····················· ···················· ···················· ····················
W 4455 hCoV-19/England/ALDP-BCA641/2020|EPI_ISL_703416|20 ···················· ···················· ····················· ···················· ···················· ····················
W 4456 hCoV-19/England/ALDP-BCAA72/2020|EPI_ISL_703418|20 ···················· ···················· ····················· ···················· ···················· ····················
W 4457 hCoV-19/England/ALDP-BCA887/2020|EPI_ISL_703420|20 ···················· ···················· ····················· ···················· ···················· ····················
W 4458 hCoV-19/England/QEUH-BC46DE/2020|EPI_ISL_703422|20 ···················· ···················· ····················· ···················· ···················· ····················
W 4459 hCoV-19/England/ALDP-BCA41A/2020|EPI_ISL_703425|20 ···················· ···················· ····················· ···················· ···················· ····················
W 4460 hCoV-19/England/QEUH-BC479F/2020|EPI_ISL_703426|20 ···················· ···················· ····················· ···················· ···················· ····················
W 4461 hCoV-19/England/QEUH-BC47EA/2020|EPI_ISL_703428|20 ···················· ···················· ····················· ···················· ···················· ····················
W 4462 hCoV-19/England/QEUH-BC4841/2020|EPI_ISL_703429|20 ···················· ···················· ····················· ···················· ···················· ····················
W 4463 hCoV-19/England/QEUH-BC453B/2020|EPI_ISL_703431|20 ···················· ···················· ····················· ···················· ···················· ····················
W 4464 hCoV-19/England/ALDP-BCA0F8/2020|EPI_ISL_703433|20 ···················· ···················· ····················· ···················· ···················· ····················
W 4465 hCoV-19/England/ALDP-BCA544/2020|EPI_ISL_703436|20 ···················· ···················· ····················· ···················· ···················· ····················
W 4466 hCoV-19/England/ALDP-BCA6D8/2020|EPI_ISL_703437|20 ···················· ···················· ····················· ···················· ···················· ····················
W 4467 hCoV-19/England/QEUH-BC42BC/2020|EPI_ISL_703439|20 ···················· ···················· ····················· ···················· ···················· ····················
W 4468 hCoV-19/England/ALDP-BCA1D7/2020|EPI_ISL_703441|20 ···················· ···················· ····················· ···················· ···················· ····················
W 4469 hCoV-19/England/QEUH-BC4920/2020|EPI_ISL_703442|20 ···················· ···················· ····················· ···················· ···················· ····················
W 4470 hCoV-19/England/QEUH-BC450E/2020|EPI_ISL_703444|20 ···················· ···················· ····················· ···················· ···················· ····················
W 4471 hCoV-19/England/QEUH-BC3ED7/2020|EPI_ISL_703445|20 ···················· ···················· ····················· ···················· ···················· ····················
W 4472 hCoV-19/England/QEUH-BC4780/2020|EPI_ISL_703447|20 ···················· ···················· ····················· ···················· ···················· ····················
W 4473 hCoV-19/England/ALDP-BCA3FF/2020|EPI_ISL_703448|20 ···················· ···················· ····················· ···················· ···················· ····················
W 4474 hCoV-19/England/ALDP-BCA6E7/2020|EPI_ISL_703450|20 ···················· ···················· ····················· ···················· ···················· ····················
W 4475 hCoV-19/England/QEUH-BC402B/2020|EPI_ISL_703453|20 ···················· ·A·················· ····················· ···················· ···················· ···A················
W 4476 hCoV-19/England/QEUH-BC48C9/2020|EPI_ISL_703456|20 ···················· ···················· ····················· ···················· ···················· ····················
W 4477 hCoV-19/England/QEUH-BC42E9/2020|EPI_ISL_703458|20 ···················· ···················· ····················· ···················· ···················· ····················
W 4478 hCoV-19/England/ALDP-BCA0DA/2020|EPI_ISL_703460|20 ···················· ···················· ····················· ···················· ···················· ····················
W 4479 hCoV-19/England/ALDP-BCA84B/2020|EPI_ISL_703461|20 ···················· ···················· ····················· ···················· ···················· ····················
W 4480 hCoV-19/England/QEUH-BC44E3/2020|EPI_ISL_703463|20 ···················· ···················· ····················· ···················· ···················· ····················
W 4481 hCoV-19/England/ALDP-BCABD9/2020|EPI_ISL_703464|20 ···················· ···················· ····················· ···················· ······T············· ····················
W 4482 hCoV-19/England/ALDP-BCA15F/2020|EPI_ISL_703467|20 ···················· ···················· ····················· ···················· ···················· ····················
W 4483 hCoV-19/England/QEUH-BC42F8/2020|EPI_ISL_703472|20 ···················· ···················· ····················· ···················· ···················· ····················
W 4484 hCoV-19/England/QEUH-BC4164/2020|EPI_ISL_703474|20 ···················· ···················· ····················· ···················· ···················· ····················
W 4485 hCoV-19/England/QEUH-BC43E6/2020|EPI_ISL_703482|20 ···················· ···················· ····················· ···················· ···················· ····················
W 4486 hCoV-19/England/ALDP-BCA78A/2020|EPI_ISL_703483|20 ···················· ···················· ····················· ···················· ···················· ····················
W 4487 hCoV-19/England/ALDP-BCA5DB/2020|EPI_ISL_703488|20 ···················· ···················· ····················· ···················· ···················· ····················
W 4488 hCoV-19/England/ALDP-BCA34A/2020|EPI_ISL_703490|20 ···················· ···················· ····················· ···················· ···················· ····················
W 4489 hCoV-19/England/QEUH-BC4823/2020|EPI_ISL_703491|20 ···················· ···················· ····················· ···················· ···················· ····················
W 4490 hCoV-19/England/ALDP-BCA08F/2020|EPI_ISL_703493|20 ···················· ···················· ····················· ···················· ···················· ····················
W 4491 hCoV-19/England/QEUH-BC3FC5/2020|EPI_ISL_703496|20 ···················· ···················· ····················· ···················· ···················· ····················
W 4492 hCoV-19/England/ALDP-BCA32C/2020|EPI_ISL_703498|20 ···················· ···················· ····················· ···················· ···················· ····················
W 4493 hCoV-19/England/QEUH-BC4595/2020|EPI_ISL_703499|20 ···················· ···················· ····················· ···················· ···················· ····················
W 4494 hCoV-19/England/CAMC-BC0B97/2020|EPI_ISL_704700|20 ···················· ···················· ····················· ···················· ···················· ····················
W 4495 hCoV-19/England/CAMC-BBFFBD/2020|EPI_ISL_704702|20 ···················· ···················· ····················· ···················· ···················· ····················
W 4496 hCoV-19/England/CAMC-BC064B/2020|EPI_ISL_704707|20 ········T··········· ···················· ····················· ···················· ···················· ····················
W 4497 hCoV-19/England/CAMC-BC0432/2020|EPI_ISL_704708|20 ···················· ···················· ····················· ···················· ···················· ····················
W 4498 hCoV-19/England/CAMC-BC0CEF/2020|EPI_ISL_704712|20 ···················· ···················· ····················· ···················· ···················· ····················
W 4499 hCoV-19/England/CAMC-BC0687/2020|EPI_ISL_704713|20 ···················· ···················· ····················· ···················· ···················· ····················
W 4500 hCoV-19/England/CAMC-BBFF62/2020|EPI_ISL_704717|20 ···················· ···················· ····················· ···················· ······T············· ····················
W 4501 hCoV-19/England/CAMC-BC011D/2020|EPI_ISL_704718|20 ···················· ···················· ····················· ···················· ···················· ····················
W 4502 hCoV-19/England/CAMC-BC0AB8/2020|EPI_ISL_704720|20 ···················· ···················· ····················· ···················· ······T············· ····················
W 4503 hCoV-19/England/CAMC-BC03BD/2020|EPI_ISL_704721|20 ···················· ···················· ····················· ···················· ···················· ····················
W 4504 hCoV-19/England/CAMC-BC0CFE/2020|EPI_ISL_704723|20 ···················· ···················· ····················· ···················· ···················· ····················
W 4505 hCoV-19/England/CAMC-BC0423/2020|EPI_ISL_704724|20 ···················· ·A·················· ····················· ···················· ···················· ···A················
W 4506 hCoV-19/England/CAMC-BC1E42/2020|EPI_ISL_704726|20 ···················· ···················· ····················· ···················· ······T············· ····················
W 4507 hCoV-19/England/CAMC-BC0AA9/2020|EPI_ISL_704727|20 ···················· ···················· ····················· ···················· ······T············· ····················
W 4508 hCoV-19/England/CAMC-BC07FD/2020|EPI_ISL_704729|20 ···················· ···················· ····················· ···················· ···················· ····················
W 4509 hCoV-19/England/CAMC-BC0186/2020|EPI_ISL_704730|20 ···················· ···················· ····················· ···················· ···················· ····················
W 4510 hCoV-19/England/CAMC-BC060F/2020|EPI_ISL_704732|20 ···················· ···················· ····················· ···················· ···················· ····················
W 4511 hCoV-19/England/CAMC-BC211B/2020|EPI_ISL_704738|20 ···················· ·A·················· ····················· ···················· ···················· ···A················
W 4512 hCoV-19/England/CAMC-BC0AD6/2020|EPI_ISL_704742|20 ···················· ···················· ····················· ···················· ······T············· ····················
W 4513 hCoV-19/England/CAMC-BC1F9A/2020|EPI_ISL_704743|20 ···················· ···················· ····················· ···················· ···················· ····················
W 4514 hCoV-19/England/CAMC-BC0AC7/2020|EPI_ISL_704745|20 ···················· ···················· ····················· ···················· ···················· ····················
W 4515 hCoV-19/England/CAMC-BBFFDB/2020|EPI_ISL_704751|20 ···················· ···················· ····················· ···················· ······T············· ····················
W 4516 hCoV-19/England/CAMC-BC19C9/2020|EPI_ISL_704753|20 ···················· ···················· ····················· ···················· ······T············· ····················
W 4517 hCoV-19/England/CAMC-BC0E70/2020|EPI_ISL_704756|20 ···················· ···················· ····················· ···················· ···················· ····················
W 4518 hCoV-19/England/CAMC-BC00A7/2020|EPI_ISL_704758|20 ···················· ·A·················· ····················· ···················· ···················· ···A················
W 4519 hCoV-19/England/CAMC-BC0CC1/2020|EPI_ISL_704761|20 ···················· ·A·················· ····················· ···················· ···················· ····················
W 4520 hCoV-19/England/CAMC-BC0CA3/2020|EPI_ISL_704764|20 ···················· ···················· ····················· ···················· ···················· ····················
W 4521 hCoV-19/England/CAMC-BC04C9/2020|EPI_ISL_704766|20 ···················· ·A·················· ····················· ···················· ···················· ···A················
W 4522 hCoV-19/England/CAMC-BC1BB4/2020|EPI_ISL_704767|20 ···················· ···················· ····················· ···················· ······T············· ····················
W 4523 hCoV-19/England/CAMC-BC0C94/2020|EPI_ISL_704769|20 ···················· ···················· ····················· ···················· ···················· ····················
W 4524 hCoV-19/England/CAMC-BC0F04/2020|EPI_ISL_704774|20 ···················· ···················· ····················· ···················· ···················· ····················
W 4525 hCoV-19/England/CAMC-BC02FC/2020|EPI_ISL_704776|20 ···················· ···················· ····················· ···················· ···················· ····················
W 4526 hCoV-19/England/CAMC-BC0001/2020|EPI_ISL_704778|20 ···················· ···················· ····················· ···················· ···················· ····················

W 4527 hCoV-19/England/CAMC-BC0C67/2020|EPI_ISL_704779|20 ···················· ···················· ····················· ···················· ···················· ···A················
W 4528 hCoV-19/England/CAMC-BC0C58/2020|EPI_ISL_704781|20 ···················· ···················· ····················· ···················· ···················· ····················
W 4529 hCoV-19/England/CAMC-BC09BB/2020|EPI_ISL_704782|20 ···················· ···················· ····················· ···················· ···················· ····················
W 4530 hCoV-19/England/CAMC-BC1C2A/2020|EPI_ISL_704785|20 ···················· ···················· ····················· ···················· ···················· ····················
W 4531 hCoV-19/England/CAMC-BC07A2/2020|EPI_ISL_704787|20 ···················· ···················· ····················· ···················· ···················· ····················
W 4532 hCoV-19/England/CAMC-BC0010/2020|EPI_ISL_704789|20 ···················· ···················· ····················· ···················· ···················· ····················
W 4533 hCoV-19/England/CAMC-BC22DC/2020|EPI_ISL_704790|20 ···················· ···················· ····················· ···················· ···················· ····················
W 4534 hCoV-19/England/CAMC-BC0EF8/2020|EPI_ISL_704792|20 ···················· ···················· ····················· ···················· ···················· ····················
W 4535 hCoV-19/England/CAMC-BC0F22/2020|EPI_ISL_704793|20 ···················· ···················· ····················· ···················· ···················· ····················
W 4536 hCoV-19/England/CAMC-BC0335/2020|EPI_ISL_704795|20 ···················· ···················· ····················· ···················· ···················· ····················
W 4537 hCoV-19/England/CAMC-BBFFAE/2020|EPI_ISL_704796|20 ···················· ···················· ····················· ···················· ···················· ····················
W 4538 hCoV-19/England/CAMC-BC2306/2020|EPI_ISL_704798|20 ···················· ···················· ····················· ···················· ···················· ····················
W 4539 hCoV-19/USA/MI-MDHHS-SC22527/2020|EPI_ISL_708230|2 ···················· ···················· ····················· ···················· ···················· ····················
W 4540 hCoV-19/USA/MI-MDHHS-SC22528/2020|EPI_ISL_708231|2 ···················· ···················· ····················· ···················· ···················· ····················
W 4541 hCoV-19/USA/MI-MDHHS-SC22408/2020|EPI_ISL_708241|2 ···················· ···················· ····················· ···················· ···················· ····················
W 4542 hCoV-19/USA/MI-MDHHS-SC22409/2020|EPI_ISL_708242|2 ···················· ···················· ····················· ···················· ···················· ····················
W 4543 hCoV-19/USA/MI-MDHHS-SC22410/2020|EPI_ISL_708243|2 ···················· ···················· ····················· ···················· ···················· ····················
W 4544 hCoV-19/USA/MI-MDHHS-SC22411/2020|EPI_ISL_708244|2 ···················· ···················· ····················· ···················· ···················· ····················
W 4545 hCoV-19/USA/MI-MDHHS-SC22412/2020|EPI_ISL_708245|2 ···················· ·············A······ ····················· ···················· ···················· ····················
W 4546 hCoV-19/USA/MI-MDHHS-SC22413/2020|EPI_ISL_708246|2 ···················· ···················· ····················· ···················· ···················· ····················
W 4547 hCoV-19/USA/MI-MDHHS-SC22414/2020|EPI_ISL_708247|2 ···················· ···················· ····················· ···················· ···················· ····················
W 4548 hCoV-19/USA/MI-MDHHS-SC22415/2020|EPI_ISL_708248|2 ···················· ···················· ····················· ···················· ···················· ····················
W 4549 hCoV-19/USA/MI-MDHHS-SC22416/2020|EPI_ISL_708249|2 ···················· ···················· ····················· ···················· ···················· ····················
W 4550 hCoV-19/USA/MI-MDHHS-SC22421/2020|EPI_ISL_708254|2 ···················· ···················· ····················· ···················· ···················· ····················
W 4551 hCoV-19/USA/MI-MDHHS-SC22422/2020|EPI_ISL_708255|2 ···················· ···················· ····················· ···················· ···················· ····················
W 4552 hCoV-19/USA/MI-MDHHS-SC22423/2020|EPI_ISL_708256|2 ···················· ···················· ····················· ···················· ···················· ····················
W 4553 hCoV-19/USA/MI-MDHHS-SC22424/2020|EPI_ISL_708257|2 ···················· ···················· ····················· ···················· ···················· ····················
W 4554 hCoV-19/USA/MI-MDHHS-SC22425/2020|EPI_ISL_708258|2 ···················· ···················· ····················· ···················· ···················· ····················
W 4555 hCoV-19/USA/MI-MDHHS-SC22426/2020|EPI_ISL_708259|2 ···················· ···················· ····················· ···················· ···················· ····················
W 4556 hCoV-19/USA/MI-MDHHS-SC22427/2020|EPI_ISL_708260|2 ···················· ···················· ····················· ···················· ···················· ····················
W 4557 hCoV-19/USA/MI-MDHHS-SC22428/2020|EPI_ISL_708261|2 ···················· ···················· ····················· ···················· ···················· ····················
W 4558 hCoV-19/USA/MI-MDHHS-SC22429/2020|EPI_ISL_708262|2 ···················· ···················· ····················· ···················· ···················· ····················
W 4559 hCoV-19/USA/MI-MDHHS-SC22430/2020|EPI_ISL_708263|2 ···················· ···················· ····················· ···················· ···················· ····················
W 4560 hCoV-19/USA/MI-MDHHS-SC22431/2020|EPI_ISL_708264|2 ···················· ···················· ····················· ···················· ···················· ····················
W 4561 hCoV-19/USA/MI-MDHHS-SC22432/2020|EPI_ISL_708265|2 ···················· ···················· ····················· ···················· ···················· ····················
W 4562 hCoV-19/USA/MI-MDHHS-SC22433/2020|EPI_ISL_708266|2 ···················· ···················· ····················· ···················· ···················· ····················
W 4563 hCoV-19/USA/MI-MDHHS-SC22434/2020|EPI_ISL_708267|2 ···················· ···················· ····················· ···················· ···················· ····················
W 4564 hCoV-19/USA/MI-MDHHS-SC22435/2020|EPI_ISL_708268|2 ···················· ···················· ····················· ···················· ···················· ····················
W 4565 hCoV-19/USA/MI-MDHHS-SC22436/2020|EPI_ISL_708269|2 ···················· ···················· ····················· ···················· ···················· ····················
W 4566 hCoV-19/USA/MI-MDHHS-SC22437/2020|EPI_ISL_708270|2 ···················· ···················· ····················· ···················· ···················· ····················
W 4567 hCoV-19/USA/MI-MDHHS-SC22438/2020|EPI_ISL_708271|2 ···················· ···················· ····················· ···················· ···················· ····················
W 4568 hCoV-19/USA/MI-MDHHS-SC22439/2020|EPI_ISL_708272|2 ···················· ···················· ····················· ···················· ···················· ····················
W 4569 hCoV-19/USA/MI-MDHHS-SC22440/2020|EPI_ISL_708273|2 ···················· ···················· ····················· ···················· ···················· ····················
W 4570 hCoV-19/USA/MI-MDHHS-SC22307/2020|EPI_ISL_708283|2 ···················· ···················· ····················· ···················· ···················· ····················
W 4571 hCoV-19/USA/MI-MDHHS-SC22308/2020|EPI_ISL_708284|2 ···················· ···················· ····················· ···················· ···················· ····················
W 4572 hCoV-19/USA/MI-MDHHS-SC22309/2020|EPI_ISL_708285|2 ···················· ·············A······ ····················· ···················· ···················· ····················
W 4573 hCoV-19/USA/MI-MDHHS-SC22310/2020|EPI_ISL_708286|2 ···················· ···················· ····················· ···················· ···················· ····················
W 4574 hCoV-19/USA/MI-MDHHS-SC22311/2020|EPI_ISL_708287|2 ···················· ·A·················· ····················· ···················· ···················· ····················
W 4575 hCoV-19/USA/MI-MDHHS-SC22312/2020|EPI_ISL_708288|2 ···················· ···················· ····················· ···················· ···················· ····················
W 4576 hCoV-19/USA/MI-MDHHS-SC22313/2020|EPI_ISL_708289|2 ···················· ···················· ····················· ···················· ···················· ····················
W 4577 hCoV-19/USA/MI-MDHHS-SC22314/2020|EPI_ISL_708290|2 ···················· ···················· ····················· ···················· ···················· ····················
W 4578 hCoV-19/USA/MI-MDHHS-SC22315/2020|EPI_ISL_708291|2 ···················· ···················· ····················· ···················· ···················· ····················
W 4579 hCoV-19/USA/MI-MDHHS-SC22316/2020|EPI_ISL_708292|2 ···················· ···················· ····················· ···················· ···················· ····················
W 4580 hCoV-19/USA/MI-MDHHS-SC22317/2020|EPI_ISL_708293|2 ···················· ···················· ····················· ···················· ···················· ····················
W 4581 hCoV-19/USA/MI-MDHHS-SC22318/2020|EPI_ISL_708294|2 ···················· ···················· ····················· ···················· ···················· ····················
W 4582 hCoV-19/USA/MI-MDHHS-SC22319/2020|EPI_ISL_708295|2 ···················· ···················· ····················· ···················· ···················· ····················
W 4583 hCoV-19/USA/MI-MDHHS-SC22320/2020|EPI_ISL_708296|2 ···················· ···················· ····················· ···················· ···················· ····················
W 4584 hCoV-19/USA/MI-MDHHS-SC22321/2020|EPI_ISL_708297|2 ···················· ···················· ····················· ···················· ···················· ····················
W 4585 hCoV-19/USA/MI-MDHHS-SC22322/2020|EPI_ISL_708298|2 ···················· ···················· ····················· ···················· ···················· ····················
W 4586 hCoV-19/USA/MI-MDHHS-SC22323/2020|EPI_ISL_708299|2 ···················· ·A·················· ····················· ···················· ···················· ····················
W 4587 hCoV-19/England/ALDP-C06833/2020|EPI_ISL_709560|20 ···················· ···················· ····················· ···················· ···················· ····················
W 4588 hCoV-19/England/QEUH-C14647/2020|EPI_ISL_709561|20 ···················· ···················· ····················· ···················· ···················· ····················
W 4589 hCoV-19/England/ALDP-C05FF3/2020|EPI_ISL_709563|20 ···················· ···················· ····················· ···················· ···················· ····················
W 4590 hCoV-19/England/QEUH-C149C6/2020|EPI_ISL_709565|20 ···················· ···················· ····················· ···················· ···················· ····················
W 4591 hCoV-19/England/MILK-BDC3FA/2020|EPI_ISL_709569|20 ···················· ···················· ····················· ···················· ···················· ···A················
W 4592 hCoV-19/England/MILK-BDB72C/2020|EPI_ISL_709570|20 ···················· ···················· ····················· ···················· ···················· ····················
W 4593 hCoV-19/England/MILK-BDBA32/2020|EPI_ISL_709572|20 ···················· ···················· ····················· ···················· ···················· ····················
W 4594 hCoV-19/England/MILK-BDB5AA/2020|EPI_ISL_709573|20 ···················· ···················· ····················· ···················· ···················· ····················
W 4595 hCoV-19/England/MILK-BDC1D2/2020|EPI_ISL_709575|20 ···················· ·A·················· ····················· ···················· ···················· ···A················
W 4596 hCoV-19/England/MILK-BDB8A1/2020|EPI_ISL_709576|20 ···················· ···················· ····················· ···················· ······T············· ····················
W 4597 hCoV-19/England/MILK-BDC309/2020|EPI_ISL_709577|20 ···················· ···················· ····················· ···················· ···················· ····················
W 4598 hCoV-19/England/MILK-BDC7A3/2020|EPI_ISL_709578|20 ···················· ···················· ····················· ···················· ···················· ····················
W 4599 hCoV-19/England/MILK-BDC5D6/2020|EPI_ISL_709579|20 ···················· ···················· ····················· ···················· ···················· ····················
W 4600 hCoV-19/England/MILK-BDC70D/2020|EPI_ISL_709585|20 ···················· ···················· ····················· ···················· ···················· ····················
W 4601 hCoV-19/England/MILK-BDC55E/2020|EPI_ISL_709588|20 ···················· ···················· ····················· ···················· ···················· ····················
W 4602 hCoV-19/England/MILK-BDB6B6/2020|EPI_ISL_709589|20 ···················· ···················· ····················· ···················· ···················· ····················
W 4603 hCoV-19/England/MILK-BDC15A/2020|EPI_ISL_709590|20 ···················· ···················· ····················· ···················· ···················· ····················
W 4604 hCoV-19/England/MILK-BDC293/2020|EPI_ISL_709591|20 ···················· ···················· ····················· ···················· ···················· ····················
W 4605 hCoV-19/England/MILK-BDBB02/2020|EPI_ISL_709592|20 ···················· ···················· ····················· ···················· ···················· ····················
W 4606 hCoV-19/England/MILK-BDCA8C/2020|EPI_ISL_709593|20 ···················· ···················· ····················· ···················· ···················· ····················

| | | | | | | | |
|---|---|---|---|---|---|---|---|
| W | 4607 hCoV-19/England/MILK-BDC9BC/2020\|EPI_ISL_709596\|20 | ···················· | ···················· | ···················· | ···················· | ···················· | ···················· |
| W | 4608 hCoV-19/England/MILK-BDC2B1/2020\|EPI_ISL_709597\|20 | ···················· | ···················· | ···················· | ···················· | ···················· | ···················· |
| W | 4609 hCoV-19/USA/AZ-TG351477/2020\|EPI_ISL_694400\|2020-0 | ···················· | ···················· | ···················· | ···················· | ···················· | ···················· |
| W | 4610 hCoV-19/USA/AZ-TG351540/2020\|EPI_ISL_694403\|2020-0 | ···················· | ···················· | ···················· | ···················· | ···················· | ···················· |
| W | 4611 hCoV-19/USA/AZ-TG351604/2020\|EPI_ISL_694404\|2020-0 | ···················· | ·A·················· | ···················· | ···················· | ···················· | ···················· |
| W | 4612 hCoV-19/USA/AZ-TG351924/2020\|EPI_ISL_694406\|2020-0 | ···················· | ···················· | ···················· | ···················· | ···················· | ···················· |
| W | 4613 hCoV-19/USA/AZ-TG352043/2020\|EPI_ISL_694422\|2020-0 | ···················· | ···················· | ···················· | ···················· | ···················· | ···················· |
| W | 4614 hCoV-19/USA/AZ-TG352045/2020\|EPI_ISL_694423\|2020-0 | ···················· | ·A·················· | ···················· | ···················· | ···················· | ···················· |
| W | 4615 hCoV-19/USA/AZ-TG352758/2020\|EPI_ISL_694428\|2020-0 | ···················· | ···················· | ···················· | ···················· | ···················· | ···················· |
| W | 4616 hCoV-19/USA/AZ-TG352877/2020\|EPI_ISL_694429\|2020-0 | ···················· | ·A·················· | ···················· | ···················· | ···················· | ···················· |
| W | 4617 hCoV-19/USA/AZ-TG352880/2020\|EPI_ISL_694430\|2020-0 | ···················· | ···················· | ···················· | ···················· | ···················· | ···················· |
| W | 4618 hCoV-19/USA/AZ-TG353013/2020\|EPI_ISL_694433\|2020-0 | ···················· | ···················· | ···················· | ···················· | ···················· | ···················· |
| W | 4619 hCoV-19/USA/AZ-TG354878/2020\|EPI_ISL_694437\|2020-0 | ···················· | ···················· | ···················· | ···················· | ···················· | ···················· |
| W | 4620 hCoV-19/USA/AZ-TG355702/2020\|EPI_ISL_694439\|2020-0 | ···················· | ···················· | ···················· | ···················· | ···················· | ···················· |
| W | 4621 hCoV-19/USA/AZ-TG357058/2020\|EPI_ISL_694440\|2020-0 | ···················· | ···················· | ···················· | ···················· | ···················· | ···················· |
| W | 4622 hCoV-19/USA/AZ-TG357125/2020\|EPI_ISL_694441\|2020-0 | ···················· | ···················· | ···················· | ···················· | ···················· | ···················· |
| W | 4623 hCoV-19/USA/AZ-TG357900/2020\|EPI_ISL_694442\|2020-0 | ···················· | ···················· | ···················· | ···················· | ···················· | ···················· |
| W | 4624 hCoV-19/USA/AZ-TG359176/2020\|EPI_ISL_694443\|2020-0 | ···················· | ·A·················· | ···················· | ···················· | ···················· | ···················· |
| W | 4625 hCoV-19/USA/AZ-TG359376/2020\|EPI_ISL_694446\|2020-0 | ···················· | ···················· | ···················· | ···················· | ···················· | ···················· |
| W | 4626 hCoV-19/USA/AZ-TG359458/2020\|EPI_ISL_694448\|2020-0 | ···················· | ···················· | ···················· | ···················· | ···················· | ···················· |
| W | 4627 hCoV-19/USA/AZ-TG359461/2020\|EPI_ISL_694449\|2020-0 | ···················· | ···················· | ···················· | ···················· | ···················· | ···················· |
| W | 4628 hCoV-19/USA/AZ-TG359582/2020\|EPI_ISL_694450\|2020-0 | ···················· | ···················· | ···················· | ···················· | ···················· | ···················· |
| W | 4629 hCoV-19/USA/AZ-TG360718/2020\|EPI_ISL_694451\|2020-0 | ···················· | ···················· | ···················· | ···················· | ···················· | ···················· |
| W | 4630 hCoV-19/USA/AZ-TG362301/2020\|EPI_ISL_694452\|2020-0 | ···················· | ···················· | ···················· | ···················· | ···················· | ···················· |
| W | 4631 hCoV-19/USA/AZ-TG363238/2020\|EPI_ISL_694453\|2020-0 | ···················· | ···················· | ···················· | ···················· | ···················· | ···················· |
| W | 4632 hCoV-19/USA/AZ-TG363241/2020\|EPI_ISL_694454\|2020-0 | ···················· | ···················· | ···················· | ···················· | ···················· | ···················· |
| W | 4633 hCoV-19/USA/AZ-TG366692/2020\|EPI_ISL_694459\|2020-0 | ···················· | ···················· | ···················· | ···················· | ···················· | ···················· |
| W | 4634 hCoV-19/USA/AZ-TG366708/2020\|EPI_ISL_694472\|2020-0 | ···················· | ·A·················· | ···················· | ···················· | ···················· | ···················· |
| W | 4635 hCoV-19/USA/AZ-TG366709/2020\|EPI_ISL_694473\|2020-0 | ···················· | ·A·················· | ···················· | ···················· | ···················· | ···················· |
| W | 4636 hCoV-19/USA/AZ-TG366710/2020\|EPI_ISL_694474\|2020-0 | ···················· | ·A·················· | ···················· | ···················· | ···················· | ···················· |
| W | 4637 hCoV-19/USA/AZ-TG366711/2020\|EPI_ISL_694475\|2020-0 | ···················· | ·A·················· | ···················· | ···················· | ···················· | ···················· |
| W | 4638 hCoV-19/USA/AZ-TG366712/2020\|EPI_ISL_694476\|2020-0 | ···················· | ·············A······ | ···················· | ···················· | ···················· | ···················· |
| W | 4639 hCoV-19/USA/AZ-TG366713/2020\|EPI_ISL_694477\|2020-0 | ···················· | ·············A······ | ···················· | ···················· | ···················· | ···················· |
| W | 4640 hCoV-19/USA/AZ-TG366716/2020\|EPI_ISL_694480\|2020-0 | ···················· | ·············A······ | ···················· | ···················· | ···················· | ···················· |
| W | 4641 hCoV-19/USA/AZ-TG366720/2020\|EPI_ISL_694484\|2020-0 | ···················· | ·A·················· | ···················· | ···················· | ···················· | ···················· |
| W | 4642 hCoV-19/USA/AZ-TG366726/2020\|EPI_ISL_694487\|2020-0 | ···················· | ···················· | ···················· | ···················· | ···················· | ···················· |
| W | 4643 hCoV-19/USA/AZ-TG366727/2020\|EPI_ISL_694488\|2020-0 | ···················· | ···················· | ···················· | ···················· | ···················· | ···················· |
| W | 4644 hCoV-19/USA/AZ-TG366731/2020\|EPI_ISL_694490\|2020-0 | ···················· | ···················· | ···················· | ···················· | ···················· | ···················· |
| W | 4645 hCoV-19/USA/AZ-TG366732/2020\|EPI_ISL_694491\|2020-0 | ···················· | ···················· | ···················· | ···················· | ···················· | ···················· |
| W | 4646 hCoV-19/USA/AZ-TG366733/2020\|EPI_ISL_694492\|2020-0 | ···················· | ·A·················· | ···················· | ···················· | ···················· | ···················· |
| W | 4647 hCoV-19/USA/AZ-TG366737/2020\|EPI_ISL_694494\|2020-0 | ···················· | ·A·················· | ···················· | ···················· | ···················· | ···················· |
| W | 4648 hCoV-19/USA/AZ-TG366740/2020\|EPI_ISL_694497\|2020-0 | ···················· | ·A·················· | ···················· | ···················· | ···················· | ···················· |
| W | 4649 hCoV-19/USA/AZ-TG366743/2020\|EPI_ISL_694499\|2020-0 | ···················· | ·A·················· | ···················· | ···················· | ···················· | ···················· |
| W | 4650 hCoV-19/USA/AZ-TG618509/2020\|EPI_ISL_695702\|2020-0 | ···················· | ·A·················· | ···················· | ···················· | ···················· | ···················· |
| W | 4651 hCoV-19/USA/AZ-TG618980/2020\|EPI_ISL_695706\|2020-0 | ···················· | ···················· | ···················· | ···················· | ···················· | ···················· |
| W | 4652 hCoV-19/USA/AZ-TG637272/2020\|EPI_ISL_695712\|2020-0 | ···················· | ·A·················· | ···················· | ···················· | ···················· | ···················· |
| W | 4653 hCoV-19/USA/AZ-TG637273/2020\|EPI_ISL_695713\|2020-0 | ···················· | ···················· | ···················· | ···················· | ···················· | ···················· |
| W | 4654 hCoV-19/USA/AZ-TG637275/2020\|EPI_ISL_695714\|2020-0 | ···················· | ···················· | ···················· | ···················· | ···················· | ···················· |
| W | 4655 hCoV-19/USA/AZ-TG637277/2020\|EPI_ISL_695715\|2020-0 | ···················· | ···················· | ···················· | ···················· | ···················· | ···················· |
| W | 4656 hCoV-19/USA/AZ-TG637281/2020\|EPI_ISL_695717\|2020-0 | ···················· | ···················· | ···················· | ···················· | ···················· | ···················· |
| W | 4657 hCoV-19/USA/AZ-TG637289/2020\|EPI_ISL_695720\|2020-0 | ···················· | ·A·················· | ···················· | ···················· | ···················· | ···················· |
| W | 4658 hCoV-19/USA/AZ-TG637304/2020\|EPI_ISL_695727\|2020-0 | ···················· | ···················· | ···················· | ···················· | ···················· | ···················· |
| W | 4659 hCoV-19/USA/AZ-TG637306/2020\|EPI_ISL_695728\|2020-0 | ···················· | ···················· | ···················· | ···················· | ···················· | ···················· |
| W | 4660 hCoV-19/USA/AZ-TG637308/2020\|EPI_ISL_695730\|2020-0 | ···················· | ···················· | ···················· | ···················· | ···················· | ···················· |
| W | 4661 hCoV-19/USA/AZ-TG637309/2020\|EPI_ISL_695731\|2020-0 | ···················· | ···················· | ···················· | ···················· | ···················· | ···················· |
| W | 4662 hCoV-19/USA/AZ-TG637315/2020\|EPI_ISL_695736\|2020-0 | ···················· | ·A·················· | ···················· | ···················· | ···················· | ···················· |
| W | 4663 hCoV-19/USA/AZ-TG637330/2020\|EPI_ISL_695740\|2020-0 | ···················· | ···················· | ···················· | ···················· | ···················· | ···················· |
| W | 4664 hCoV-19/USA/AZ-TG637333/2020\|EPI_ISL_695742\|2020-0 | ···················· | ···················· | ···················· | ···················· | ···················· | ···················· |
| W | 4665 hCoV-19/USA/AZ-TG637339/2020\|EPI_ISL_695744\|2020-0 | ···················· | ···················· | ···················· | ···················· | ···················· | ···················· |
| W | 4666 hCoV-19/USA/AZ-TG637346/2020\|EPI_ISL_695745\|2020-0 | ···················· | ···················· | ···················· | ···················· | ···················· | ···················· |
| W | 4667 hCoV-19/USA/AZ-TG637356/2020\|EPI_ISL_695749\|2020-0 | ···················· | ·A·················· | ···················· | ···················· | ···················· | ···················· |
| W | 4668 hCoV-19/USA/AZ-TG640780/2020\|EPI_ISL_695755\|2020-0 | ···················· | ···················· | ···················· | ···················· | ···················· | ···················· |
| W | 4669 hCoV-19/USA/AZ-TG640781/2020\|EPI_ISL_695756\|2020-0 | ···················· | ···················· | ···················· | ···················· | ···················· | ···················· |
| W | 4670 hCoV-19/USA/AZ-TG640783/2020\|EPI_ISL_695757\|2020-0 | ···················· | ···················· | ···················· | ···················· | ···················· | ···················· |
| W | 4671 hCoV-19/USA/AZ-TG640786/2020\|EPI_ISL_695759\|2020-0 | ···················· | ···················· | ···················· | ···················· | ···················· | ···················· |
| W | 4672 hCoV-19/USA/AZ-TG640787/2020\|EPI_ISL_695760\|2020-0 | ···················· | ···················· | ···················· | ···················· | ···················· | ···················· |
| W | 4673 hCoV-19/USA/AZ-TG640790/2020\|EPI_ISL_695762\|2020-0 | ···················· | ···················· | ···················· | ···················· | ···················· | ···················· |
| W | 4674 hCoV-19/USA/AZ-TG640791/2020\|EPI_ISL_695763\|2020-0 | ···················· | ···················· | ···················· | ···················· | ···················· | ···················· |
| W | 4675 hCoV-19/USA/AZ-TG640793/2020\|EPI_ISL_695764\|2020-0 | ···················· | ···················· | ···················· | ···················· | ···················· | ···················· |
| W | 4676 hCoV-19/USA/AZ-TG640794/2020\|EPI_ISL_695765\|2020-0 | ···················· | ···················· | ···················· | ···················· | ···················· | ···················· |
| W | 4677 hCoV-19/USA/AZ-TG640795/2020\|EPI_ISL_695766\|2020-0 | ···················· | ···················· | ···················· | ···················· | ···················· | ···················· |
| W | 4678 hCoV-19/USA/AZ-TG640797/2020\|EPI_ISL_695768\|2020-0 | ···················· | ···················· | ···················· | ···················· | ···················· | ···················· |
| W | 4679 hCoV-19/USA/AZ-TG640815/2020\|EPI_ISL_695772\|2020-0 | ···················· | ···················· | ···················· | ···················· | ···················· | ···················· |
| W | 4680 hCoV-19/USA/AZ-TG640816/2020\|EPI_ISL_695773\|2020-0 | ···················· | ···················· | ···················· | ···················· | ···················· | ···················· |
| W | 4681 hCoV-19/USA/AZ-TG640818/2020\|EPI_ISL_695774\|2020-0 | ···················· | ···················· | ···················· | ···················· | ···················· | ···················· |
| W | 4682 hCoV-19/USA/AZ-TG640819/2020\|EPI_ISL_695775\|2020-0 | ···················· | ···················· | ···················· | ···················· | ···················· | ···················· |
| W | 4683 hCoV-19/USA/AZ-TG640821/2020\|EPI_ISL_695777\|2020-0 | ···················· | ···················· | ···················· | ···················· | ···················· | ···················· |
| W | 4684 hCoV-19/USA/AZ-TG640822/2020\|EPI_ISL_695778\|2020-0 | ···················· | ·A·················· | ···················· | ···················· | ···················· | ···················· |
| W | 4685 hCoV-19/USA/AZ-TG640826/2020\|EPI_ISL_695781\|2020-0 | ···················· | ···················· | ···················· | ···················· | ···················· | ···················· |
| W | 4686 hCoV-19/USA/AZ-TG640827/2020\|EPI_ISL_695782\|2020-0 | ···················· | ···················· | ···················· | ···················· | ···················· | ···················· |

| | | | | | | | |
|---|---|---|---|---|---|---|---|
| W | 4687 hCoV-19/USA/AZ-TG640830/2020\|EPI_ISL_695784\|2020-0 | ···················· | ···················· | ····················· | ···················· | ···················· | ···················· |
| W | 4688 hCoV-19/USA/AZ-TG640839/2020\|EPI_ISL_695787\|2020-0 | ···················· | ···················· | ····················· | ···················· | ···················· | ···················· |
| W | 4689 hCoV-19/USA/AZ-TG640843/2020\|EPI_ISL_695788\|2020-0 | ···················· | ···················· | ····················· | ···················· | ···················· | ···················· |
| W | 4690 hCoV-19/USA/AZ-TG640844/2020\|EPI_ISL_695789\|2020-0 | ···················· | ···················· | ····················· | ···················· | ···················· | ···················· |
| W | 4691 hCoV-19/USA/AZ-TG640845/2020\|EPI_ISL_695790\|2020-0 | ···················· | ···················· | ····················· | ···················· | ···················· | ···················· |
| W | 4692 hCoV-19/USA/AZ-TG640851/2020\|EPI_ISL_695794\|2020-0 | ···················· | ···················· | ····················· | ···················· | ···················· | ···················· |
| W | 4693 hCoV-19/USA/AZ-TG640852/2020\|EPI_ISL_695795\|2020-0 | ···················· | ···················· | ····················· | ···················· | ···················· | ···················· |
| W | 4694 hCoV-19/USA/AZ-TG640853/2020\|EPI_ISL_695796\|2020-0 | ···················· | ···················· | ····················· | ···················· | ···················· | ···················· |
| W | 4695 hCoV-19/USA/AZ-TG640854/2020\|EPI_ISL_695797\|2020-0 | ···················· | ···················· | ····················· | ···················· | ···················· | ···················· |
| W | 4696 hCoV-19/USA/AZ-TG640858/2020\|EPI_ISL_695798\|2020-0 | ···················· | ···················· | ····················· | ···················· | ···················· | ···················· |
| W | 4697 hCoV-19/England/CAMC-BD1E7C/2020\|EPI_ISL_702144\|20 | ···················· | ···················· | ····················· | ···················· | ···················· | ···················· |
| W | 4698 hCoV-19/England/CAMC-BD1E30/2020\|EPI_ISL_702145\|20 | ···················· | ···················· | ····················· | ···················· | ···················· | ···················· |
| W | 4699 hCoV-19/England/CAMC-BD146B/2020\|EPI_ISL_702147\|20 | ···················· | ···················· | ····················· | ···················· | ···················· | ···················· |
| W | 4700 hCoV-19/England/CAMC-BD1322/2020\|EPI_ISL_702148\|20 | ···················· | ···················· | ····················· | ···················· | ···················· | ···················· |
| W | 4701 hCoV-19/England/CAMC-BD488A/2020\|EPI_ISL_702149\|20 | ···················· | ···················· | ····················· | ···················· | ···················· | ···················· |
| W | 4702 hCoV-19/England/CAMC-BD487B/2020\|EPI_ISL_702171\|20 | ···················· | ···················· | ····················· | ···················· | ·····T·············· | ···················· |
| W | 4703 hCoV-19/England/CAMC-BD18AB/2020\|EPI_ISL_702172\|20 | ···················· | ···················· | ····················· | ···················· | ···················· | ···················· |
| W | 4704 hCoV-19/England/CAMC-BD2AE0/2020\|EPI_ISL_702173\|20 | ···················· | ···················· | ····················· | ···················· | ···················· | ···················· |
| W | 4705 hCoV-19/England/CAMC-BD18BA/2020\|EPI_ISL_702179\|20 | ···················· | ···················· | ····················· | ···················· | ···················· | ···················· |
| W | 4706 hCoV-19/England/CAMC-BD2118/2020\|EPI_ISL_702184\|20 | ···················· | ···················· | ····················· | ···················· | ···················· | ···················· |
| W | 4707 hCoV-19/England/CAMC-BD2BFC/2020\|EPI_ISL_702187\|20 | ···················· | ···················· | ····················· | ···················· | ···················· | ···················· |
| W | 4708 hCoV-19/England/CAMC-BD1DF7/2020\|EPI_ISL_702188\|20 | ···················· | ···················· | ····················· | ···················· | ···················· | ···················· |
| W | 4709 hCoV-19/England/CAMC-BD43E3/2020\|EPI_ISL_702189\|20 | ···················· | ···················· | ····················· | ···················· | ·····T·············· | ···················· |
| W | 4710 hCoV-19/England/CAMC-BD29F2/2020\|EPI_ISL_702195\|20 | ···················· | ···················· | ····················· | ···················· | ···················· | ···················· |
| W | 4711 hCoV-19/England/CAMC-BD3A94/2020\|EPI_ISL_702196\|20 | ···················· | ···················· | ····················· | ···················· | ···················· | ···················· |
| W | 4712 hCoV-19/England/CAMC-BD2B65/2020\|EPI_ISL_702198\|20 | ···················· | ···················· | ····················· | ···················· | ···················· | ···················· |
| W | 4713 hCoV-19/England/CAMC-BD1A2D/2020\|EPI_ISL_702201\|20 | ···················· | ···················· | ····················· | ···················· | ···················· | ···················· |
| W | 4714 hCoV-19/England/CAMC-BD137D/2020\|EPI_ISL_702202\|20 | ···················· | ···················· | ····················· | ···················· | ···················· | ···················· |
| W | 4715 hCoV-19/England/CAMC-BD1BDF/2020\|EPI_ISL_702203\|20 | ···················· | ···················· | ····················· | ···················· | ···················· | ···················· |
| W | 4716 hCoV-19/England/CAMC-BD17BD/2020\|EPI_ISL_702206\|20 | ···················· | ···················· | ····················· | ···················· | ···················· | ···················· |
| W | 4717 hCoV-19/England/ALDP-BCA67E/2020\|EPI_ISL_703475\|20 | ···················· | ···················· | ····················· | ···················· | ···················· | ···················· |
| W | 4718 hCoV-19/England/ALDP-BCA2A7/2020\|EPI_ISL_703477\|20 | ···················· | ···················· | ····················· | ···················· | ···················· | ···················· |
| W | 4719 hCoV-19/England/ALDP-BCA1AA/2020\|EPI_ISL_703478\|20 | ···················· | ···················· | ····················· | ···················· | ···················· | ···················· |
| W | 4720 hCoV-19/England/QEUH-BC41A0/2020\|EPI_ISL_703501\|20 | ···················· | ···················· | ····················· | ···················· | ···················· | ···················· |
| W | 4721 hCoV-19/England/ALDP-BCA9B1/2020\|EPI_ISL_703503\|20 | ···················· | ···················· | ····················· | ···················· | ···················· | ···················· |
| W | 4722 hCoV-19/England/ALDP-BCA9A2/2020\|EPI_ISL_703504\|20 | ···················· | ···················· | ····················· | ···················· | ···················· | ···················· |
| W | 4723 hCoV-19/England/QEUH-BC3F7A/2020\|EPI_ISL_703506\|20 | ···················· | ···················· | ····················· | ···················· | ···················· | ···················· |
| W | 4724 hCoV-19/England/ALDP-BCAAAF/2020\|EPI_ISL_703507\|20 | ···················· | ···················· | ····················· | ···················· | ···················· | ···················· |
| W | 4725 hCoV-19/England/ALDP-BCA4B0/2020\|EPI_ISL_703509\|20 | ···················· | ···················· | ····················· | ···················· | ···················· | ···················· |
| W | 4726 hCoV-19/England/QEUH-BC4410/2020\|EPI_ISL_703510\|20 | ···················· | ···················· | ····················· | ···················· | ···················· | ···················· |
| W | 4727 hCoV-19/England/ALDP-BCA82D/2020\|EPI_ISL_703518\|20 | ···················· | ···················· | ····················· | ···················· | ···················· | ···················· |
| W | 4728 hCoV-19/England/ALDP-BCA4ED/2020\|EPI_ISL_703520\|20 | ···················· | ···················· | ····················· | ···················· | ···················· | ···················· |
| W | 4729 hCoV-19/England/QEUH-BC489C/2020\|EPI_ISL_703522\|20 | ···················· | ···················· | ····················· | ···················· | ···················· | ···················· |
| W | 4730 hCoV-19/England/QEUH-BC403A/2020\|EPI_ISL_703525\|20 | ···················· | ···················· | ····················· | ···················· | ···················· | ···················· |
| W | 4731 hCoV-19/England/ALDP-BCA483/2020\|EPI_ISL_703526\|20 | ···················· | ···················· | ····················· | ···················· | ···················· | ···················· |
| W | 4732 hCoV-19/England/ALDP-BCABE8/2020\|EPI_ISL_703528\|20 | ···················· | ···················· | ····················· | ···················· | ···················· | ···················· |
| W | 4733 hCoV-19/England/QEUH-BC4902/2020\|EPI_ISL_703529\|20 | ···················· | ···················· | ····················· | ···················· | ···················· | ···················· |
| W | 4734 hCoV-19/England/ALDP-BCA3B3/2020\|EPI_ISL_703531\|20 | ···················· | ···················· | ····················· | ···················· | ···················· | ···················· |
| W | 4735 hCoV-19/England/ALDP-BCA957/2020\|EPI_ISL_703532\|20 | ···················· | ···················· | ····················· | ···················· | ···················· | ···················· |
| W | 4736 hCoV-19/England/QEUH-BC48D8/2020\|EPI_ISL_703534\|20 | ···················· | ···················· | ····················· | ···················· | ···················· | ···················· |
| W | 4737 hCoV-19/England/ALDP-BCA034/2020\|EPI_ISL_703536\|20 | ···················· | ···················· | ····················· | ···················· | ···················· | ···················· |
| W | 4738 hCoV-19/England/QEUH-BC4638/2020\|EPI_ISL_703538\|20 | ···················· | ···················· | ····················· | ···················· | ···················· | ···················· |
| W | 4739 hCoV-19/England/ALDP-BCA2D4/2020\|EPI_ISL_703539\|20 | ···················· | ···················· | ····················· | ···················· | ···················· | ···················· |
| W | 4740 hCoV-19/England/CAMC-BC049C/2020\|EPI_ISL_704800\|20 | ···················· | ···················· | ····················· | ···················· | ···················· | ···················· |
| W | 4741 hCoV-19/England/CAMC-BC0854/2020\|EPI_ISL_704801\|20 | ···················· | ·A·················· | ····················· | ···················· | ···················· | ···················· |
| W | 4742 hCoV-19/England/CAMC-BC0229/2020\|EPI_ISL_704803\|20 | ···················· | ···················· | ····················· | ···················· | ···················· | ···················· |
| W | 4743 hCoV-19/England/CAMC-BC03DB/2020\|EPI_ISL_704805\|20 | ···················· | ···················· | ····················· | ···················· | ···················· | ···················· |
| W | 4744 hCoV-19/England/CAMC-BBFFF9/2020\|EPI_ISL_704808\|20 | ···················· | ···················· | ····················· | ···················· | ···················· | ···················· |
| W | 4745 hCoV-19/England/CAMC-BC0942/2020\|EPI_ISL_704810\|20 | ···················· | ···················· | ····················· | ···················· | ···················· | ···················· |
| W | 4746 hCoV-19/England/CAMC-BC0CB2/2020\|EPI_ISL_704811\|20 | ···················· | ···················· | ····················· | ···················· | ···················· | ···················· |
| W | 4747 hCoV-19/England/MILK-A91C7F/2020\|EPI_ISL_704812\|20 | ···················· | ···················· | ····················· | ···················· | ···················· | ···················· |
| W | 4748 hCoV-19/England/CAMC-BC1BC3/2020\|EPI_ISL_704813\|20 | ···················· | ···················· | ····················· | ···················· | ···················· | ···················· |
| W | 4749 hCoV-19/England/CAMC-BC05C6/2020\|EPI_ISL_704814\|20 | ···················· | ···················· | ····················· | ···················· | ···················· | ···················· |
| W | 4750 hCoV-19/England/MILK-B3CFE5/2020\|EPI_ISL_704815\|20 | ···················· | ···················· | ····················· | ···················· | ···················· | ···················· |
| W | 4751 hCoV-19/England/MILK-B3C738/2020\|EPI_ISL_704817\|20 | ···················· | ···················· | ····················· | ···················· | ···················· | ···················· |
| W | 4752 hCoV-19/England/CAMC-BC071B/2020\|EPI_ISL_704819\|20 | ···················· | ···················· | ····················· | ···················· | ···················· | ···················· |
| W | 4753 hCoV-19/England/CAMC-BC0089/2020\|EPI_ISL_704821\|20 | ···················· | ···················· | ····················· | ···················· | ···················· | ···················· |
| W | 4754 hCoV-19/England/CAMC-BC0818/2020\|EPI_ISL_704824\|20 | ···················· | ···················· | ····················· | ···················· | ···················· | ···················· |
| W | 4755 hCoV-19/England/CAMC-BC055D/2020\|EPI_ISL_704825\|20 | ···················· | ······A············· | ····················· | ···················· | ···················· | ···················· |
| W | 4756 hCoV-19/England/CAMC-BC0E61/2020\|EPI_ISL_704827\|20 | ···················· | ···················· | ····················· | ···················· | ·····T·············· | ···················· |
| W | 4757 hCoV-19/England/CAMC-BBFF17/2020\|EPI_ISL_704830\|20 | ···················· | ···················· | ····················· | ···················· | ···················· | ···················· |
| W | 4758 hCoV-19/England/CAMC-BBFECF/2020\|EPI_ISL_704833\|20 | ···················· | ·A·················· | ····················· | ···················· | ···················· | ···················· |
| W | 4759 hCoV-19/England/CAMC-BC0DFB/2020\|EPI_ISL_704837\|20 | ···················· | ···················· | ····················· | ···················· | ···················· | ···················· |
| W | 4760 hCoV-19/England/CAMC-BC0450/2020\|EPI_ISL_704840\|20 | ···················· | ···················· | ····················· | ···················· | ···················· | ···················· |
| W | 4761 hCoV-19/England/CAMC-BC1E51/2020\|EPI_ISL_704841\|20 | ···················· | ···················· | ····················· | ···················· | ·····T·············· | ···················· |
| W | 4762 hCoV-19/England/CAMC-BBC2C6/2020\|EPI_ISL_704842\|20 | ···················· | ···················· | ····················· | ···················· | ···················· | ···················· |
| W | 4763 hCoV-19/England/CAMC-BC0274/2020\|EPI_ISL_704844\|20 | ···················· | ···················· | ····················· | ···················· | ···················· | ···················· |
| W | 4764 hCoV-19/England/CAMC-BC08BE/2020\|EPI_ISL_704849\|20 | ···················· | ···················· | ····················· | ···················· | ···················· | ···················· |
| W | 4765 hCoV-19/England/NORW-F654A/2020\|EPI_ISL_704850\|202 | ···················· | ···················· | ····················· | ···················· | ···················· | ···················· |
| W | 4766 hCoV-19/England/CAMC-BC0E25/2020\|EPI_ISL_704851\|20 | ···················· | ···················· | ····················· | ···················· | ···················· | ···················· |

| | | | | | | | |
|---|---|---|---|---|---|---|---|
| W | 4767 hCoV-19/England/CAMC-BC1EF7/2020\|EPI_ISL_704852\|20 | ···················· | ···················· | ···················· | ···················· | ······T············· | ···················· |
| W | 4768 hCoV-19/England/NORW-F6C54/2020\|EPI_ISL_704853\|202 | ···················· | ···················· | ···················· | ···················· | ···················· | ···················· |
| W | 4769 hCoV-19/England/CAMC-BC0ECB/2020\|EPI_ISL_704854\|20 | ···················· | ···················· | ···················· | ···················· | ···················· | ···················· |
| W | 4770 hCoV-19/England/NORW-F7637/2020\|EPI_ISL_704855\|202 | ···················· | ···················· | ···················· | ···················· | ···················· | ···················· |
| W | 4771 hCoV-19/England/CAMC-BC04AB/2020\|EPI_ISL_704857\|20 | ···················· | ···················· | ···················· | ···················· | ···················· | ···················· |
| W | 4773 hCoV-19/England/CAMC-BC01C2/2020\|EPI_ISL_704859\|20 | ···················· | ···················· | ···················· | ···················· | ···················· | ···················· |
| W | 4774 hCoV-19/England/CAMC-BBFF44/2020\|EPI_ISL_704860\|20 | ···················· | ···················· | ···················· | ···················· | ···················· | ···················· |
| W | 4775 hCoV-19/England/MILK-B3B13B/2020\|EPI_ISL_704861\|20 | ···················· | ···················· | ···················· | ···················· | ···················· | ···················· |
| W | 4776 hCoV-19/England/CAMC-BC0F9B/2020\|EPI_ISL_704863\|20 | ···················· | ···················· | ···················· | ···················· | ···················· | ···················· |
| W | 4777 hCoV-19/England/MILK-B3B1D1/2020\|EPI_ISL_704864\|20 | ···················· | ···················· | ···················· | ···················· | ···················· | ···················· |
| W | 4778 hCoV-19/England/CAMC-BC0836/2020\|EPI_ISL_704865\|20 | ···················· | ···················· | ···················· | ···················· | ···················· | ···················· |
| W | 4779 hCoV-19/England/MILK-A916DB/2020\|EPI_ISL_704866\|20 | ···················· | ·A·················· | ···················· | ···················· | ···················· | ···A················ |
| W | 4780 hCoV-19/England/CAMC-BC1BE1/2020\|EPI_ISL_704868\|20 | ···················· | ···················· | ···················· | ···················· | ······T············· | ···················· |
| W | 4781 hCoV-19/England/MILK-A91AEE/2020\|EPI_ISL_704869\|20 | ···················· | ·A·················· | ···················· | ···················· | ···················· | ···A················ |
| W | 4782 hCoV-19/England/SHEF-CAFF6/2020\|EPI_ISL_707000\|202 | ···················· | ···················· | ···················· | ···················· | ···················· | ···················· |
| W | 4783 hCoV-19/England/SHEF-CCD27/2020\|EPI_ISL_707001\|202 | ···················· | ···················· | ···················· | ···················· | ······T············· | ···················· |
| W | 4784 hCoV-19/England/SHEF-CDA5C/2020\|EPI_ISL_707002\|202 | ···················· | ···················· | ···················· | ···················· | ···················· | ···················· |
| W | 4785 hCoV-19/England/SHEF-CE402/2020\|EPI_ISL_707003\|202 | ···················· | ···················· | ···················· | ···················· | ···················· | ···················· |
| W | 4786 hCoV-19/England/SHEF-CE99A/2020\|EPI_ISL_707004\|202 | ···················· | ·A·················· | ···················· | ···················· | ···················· | ···A················ |
| W | 4787 hCoV-19/England/SHEF-D0E78/2020\|EPI_ISL_707005\|202 | ···················· | ···················· | ···················· | ···················· | ···················· | ···················· |
| W | 4788 hCoV-19/England/SHEF-D2C12/2020\|EPI_ISL_707006\|202 | ···················· | ···················· | ···················· | ···················· | ···················· | ···················· |
| W | 4789 hCoV-19/USA/MI-MDHHS-SC22324/2020\|EPI_ISL_708300\|2 | ···················· | ·A·················· | ···················· | ···················· | ···················· | ···················· |
| W | 4790 hCoV-19/USA/MI-MDHHS-SC22325/2020\|EPI_ISL_708301\|2 | ···················· | ···················· | ···················· | ···················· | ···················· | ···················· |
| W | 4791 hCoV-19/USA/MI-MDHHS-SC22326/2020\|EPI_ISL_708302\|2 | ···················· | ···················· | ···················· | ···················· | ···················· | ···················· |
| W | 4792 hCoV-19/USA/MI-MDHHS-SC22327/2020\|EPI_ISL_708303\|2 | ···················· | ·A·················· | ···················· | ···················· | ···················· | ···················· |
| W | 4793 hCoV-19/USA/MI-MDHHS-SC22328/2020\|EPI_ISL_708304\|2 | ···················· | ···················· | ···················· | ···················· | ···················· | ···················· |
| W | 4794 hCoV-19/USA/MI-MDHHS-SC22329/2020\|EPI_ISL_708305\|2 | ···················· | ······A······A······ | ···················· | ···················· | ···················· | ···················· |
| W | 4795 hCoV-19/Switzerland/GE-HUG-54280/2020\|EPI_ISL_7083 | ···················· | ···················· | ···················· | ···················· | ···················· | ···················· |
| W | 4796 hCoV-19/Switzerland/GE-HUG-57410/2020\|EPI_ISL_7083 | ···················· | ···················· | ···················· | ···················· | ···················· | ···················· |
| W | 4797 hCoV-19/Switzerland/GE-HUG-88642/2020\|EPI_ISL_7083 | ···················· | ···················· | ···················· | ···················· | ···················· | ···················· |
| W | 4798 hCoV-19/Switzerland/GE-HUG-18268/2020\|EPI_ISL_7083 | ···················· | ···················· | ···················· | ···················· | ···················· | ···················· |
| W | 4799 hCoV-19/USA/MI-MDHHS-SC22374/2020\|EPI_ISL_708354\|2 | ···················· | ···················· | ···················· | ···················· | ···················· | ···················· |
| W | 4800 hCoV-19/USA/MI-MDHHS-SC22375/2020\|EPI_ISL_708355\|2 | ···················· | ···················· | ···················· | ···················· | ···················· | ···················· |
| W | 4801 hCoV-19/USA/MI-MDHHS-SC22376/2020\|EPI_ISL_708356\|2 | ···················· | ·A·················· | ···················· | ···················· | ···················· | ···················· |
| W | 4802 hCoV-19/USA/MI-MDHHS-SC22377/2020\|EPI_ISL_708357\|2 | ···················· | ···················· | ···················· | ···················· | ···················· | ···················· |
| W | 4803 hCoV-19/USA/MI-MDHHS-SC22378/2020\|EPI_ISL_708358\|2 | ···················· | ·············A······ | ···················· | ···················· | ···················· | ···················· |
| W | 4804 hCoV-19/USA/MI-MDHHS-SC22380/2020\|EPI_ISL_708360\|2 | ···················· | ···················· | ···················· | ···················· | ···················· | ···················· |
| W | 4805 hCoV-19/USA/MI-MDHHS-SC22381/2020\|EPI_ISL_708361\|2 | ···················· | ·············A······ | ···················· | ···················· | ···················· | ···················· |
| W | 4806 hCoV-19/USA/MI-MDHHS-SC22382/2020\|EPI_ISL_708362\|2 | ···················· | ·············A······ | ···················· | ···················· | ···················· | ···················· |
| W | 4807 hCoV-19/USA/MI-MDHHS-SC22383/2020\|EPI_ISL_708363\|2 | ···················· | ···················· | ···················· | ···················· | ···················· | ···················· |
| W | 4808 hCoV-19/USA/MI-MDHHS-SC22384/2020\|EPI_ISL_708364\|2 | ···················· | ···················· | ···················· | ···················· | ···················· | ···················· |
| W | 4809 hCoV-19/USA/MI-MDHHS-SC22385/2020\|EPI_ISL_708365\|2 | ···················· | ···················· | ···················· | ···················· | ···················· | ···················· |
| W | 4810 hCoV-19/USA/MI-MDHHS-SC22386/2020\|EPI_ISL_708366\|2 | ···················· | ···················· | ···················· | ···················· | ···················· | ···················· |
| W | 4811 hCoV-19/USA/MI-MDHHS-SC22387/2020\|EPI_ISL_708367\|2 | ···················· | ·············A······ | ···················· | ···················· | ···················· | ···················· |
| W | 4812 hCoV-19/USA/MI-MDHHS-SC22388/2020\|EPI_ISL_708368\|2 | ···················· | ···················· | ···················· | ···················· | ···················· | ···················· |
| W | 4813 hCoV-19/USA/MI-MDHHS-SC22389/2020\|EPI_ISL_708369\|2 | ···················· | ···················· | ···················· | ···················· | ···················· | ···················· |
| W | 4814 hCoV-19/USA/MI-MDHHS-SC22390/2020\|EPI_ISL_708370\|2 | ···················· | ···················· | ···················· | ···················· | ···················· | ···················· |
| W | 4815 hCoV-19/USA/MI-MDHHS-SC22391/2020\|EPI_ISL_708371\|2 | ···················· | ···················· | ···················· | ···················· | ···················· | ···················· |
| W | 4816 hCoV-19/USA/MI-MDHHS-SC22392/2020\|EPI_ISL_708372\|2 | ···················· | ···················· | ···················· | ···················· | ···················· | ···················· |
| W | 4817 hCoV-19/USA/MI-MDHHS-SC22393/2020\|EPI_ISL_708373\|2 | ···················· | ···················· | ···················· | ···················· | ···················· | ···················· |
| W | 4818 hCoV-19/USA/MI-MDHHS-SC22394/2020\|EPI_ISL_708374\|2 | ···················· | ···················· | ···················· | ···················· | ···················· | ···················· |
| W | 4819 hCoV-19/USA/MI-MDHHS-SC22395/2020\|EPI_ISL_708375\|2 | ···················· | ···················· | ···················· | ···················· | ···················· | ···················· |
| W | 4820 hCoV-19/USA/MI-MDHHS-SC22396/2020\|EPI_ISL_708376\|2 | ···················· | ···················· | ···················· | ···················· | ···················· | ···················· |
| W | 4821 hCoV-19/USA/MI-MDHHS-SC22397/2020\|EPI_ISL_708377\|2 | ···················· | ···················· | ···················· | ···················· | ···················· | ···················· |
| W | 4822 hCoV-19/USA/MI-MDHHS-SC22398/2020\|EPI_ISL_708378\|2 | ···················· | ···················· | ···················· | ···················· | ···················· | ···················· |
| W | 4823 hCoV-19/Switzerland/GE-HUG-40501/2020\|EPI_ISL_7083 | ···················· | ···················· | ···················· | ···················· | ···················· | ···················· |
| W | 4824 hCoV-19/England/MILK-BDC5C7/2020\|EPI_ISL_709605\|20 | ···················· | ···················· | ···················· | ···················· | ···················· | ···················· |
| W | 4825 hCoV-19/England/MILK-BDC318/2020\|EPI_ISL_709608\|20 | ···················· | ···················· | ···················· | ···················· | ···················· | ···················· |
| W | 4826 hCoV-19/England/MILK-BDC2EE/2020\|EPI_ISL_709609\|20 | ···················· | ···················· | ···················· | ···················· | ···················· | ···················· |
| W | 4827 hCoV-19/England/MILK-BDB8CF/2020\|EPI_ISL_709614\|20 | ···················· | ···················· | ···················· | ···················· | ···················· | ···················· |
| W | 4828 hCoV-19/England/MILK-BDB926/2020\|EPI_ISL_709615\|20 | ···················· | ···················· | ···················· | ···················· | ···················· | ···················· |
| W | 4829 hCoV-19/England/MILK-BDC679/2020\|EPI_ISL_709618\|20 | ···················· | ···················· | ···················· | ···················· | ···················· | ···················· |
| W | 4830 hCoV-19/England/MILK-BDC0D5/2020\|EPI_ISL_709619\|20 | ···················· | ···················· | ···················· | ···················· | ···················· | ···················· |
| W | 4831 hCoV-19/England/MILK-BDBA8D/2020\|EPI_ISL_709623\|20 | ···················· | ···················· | ···················· | ···················· | ···················· | ···················· |
| W | 4832 hCoV-19/England/MILK-BDC767/2020\|EPI_ISL_709625\|20 | ···················· | ···················· | ···················· | ···················· | ···················· | ···················· |
| W | 4833 hCoV-19/England/MILK-BDB795/2020\|EPI_ISL_709629\|20 | ···················· | ···················· | ···················· | ···················· | ···················· | ···················· |
| W | 4834 hCoV-19/England/MILK-BDB6C5/2020\|EPI_ISL_709631\|20 | ···················· | ···················· | ···················· | ···················· | ···················· | ···················· |
| W | 4835 hCoV-19/England/MILK-BDB5B9/2020\|EPI_ISL_709632\|20 | ···················· | ···················· | ···················· | ···················· | ···················· | ···················· |
| W | 4836 hCoV-19/England/MILK-BDBA14/2020\|EPI_ISL_709637\|20 | ···················· | ···················· | ···················· | ···················· | ···················· | ···················· |
| W | 4837 hCoV-19/England/MILK-BDCAAA/2020\|EPI_ISL_709640\|20 | ···················· | ···················· | ···················· | ···················· | ···················· | ···················· |
| W | 4838 hCoV-19/England/MILK-BDB64D/2020\|EPI_ISL_709644\|20 | ···················· | ·A·················· | ···················· | ···················· | ···················· | ···A················ |
| W | 4839 hCoV-19/England/MILK-BDB58C/2020\|EPI_ISL_709646\|20 | ···················· | ···················· | ···················· | ···················· | ···················· | ···················· |
| W | 4840 hCoV-19/England/MILK-BDB9EA/2020\|EPI_ISL_709647\|20 | ···················· | ···················· | ···················· | ···················· | ···················· | ···················· |
| W | 4841 hCoV-19/England/MILK-BDB8ED/2020\|EPI_ISL_709648\|20 | ···················· | ···················· | ···················· | ···················· | ···················· | ···················· |
| W | 4842 hCoV-19/England/MILK-BDC9CB/2020\|EPI_ISL_709650\|20 | ···················· | ···················· | ···················· | ···················· | ···················· | ···················· |
| W | 4843 hCoV-19/England/MILK-BDCAE6/2020\|EPI_ISL_709652\|20 | ···················· | ···················· | ···················· | ···················· | ···················· | ···················· |
| W | 4844 hCoV-19/England/MILK-BDC2DF/2020\|EPI_ISL_709654\|20 | ···················· | ···················· | ···················· | ···················· | ······T············· | ···················· |
| W | 4845 hCoV-19/England/MILK-BDB4E9/2020\|EPI_ISL_709655\|20 | ···················· | ···················· | ···················· | ···················· | ···················· | ···················· |
| W | 4846 hCoV-19/England/MILK-BDB6E3/2020\|EPI_ISL_709656\|20 | ···················· | ···················· | ···················· | ···················· | ···················· | ···················· |
| W | 4847 hCoV-19/England/MILK-BDC7B2/2020\|EPI_ISL_709658\|20 | ···················· | ···················· | ···················· | ···················· | ···················· | ···················· |

| | | | | | | | | |
|---|---|---|---|---|---|---|---|---|
| W | 4848 hCoV-19/England/MILK-BDBF8E/2020\|EPI_ISL_709659\|20 | ···················· | ···················· | ···················· | ···················· | ···················· | ···················· | |
| W | 4849 hCoV-19/England/MILK-BDC72B/2020\|EPI_ISL_709662\|20 | ···················· | ···················· | ···················· | ···················· | ···················· | ···················· | |
| W | 4850 hCoV-19/England/MILK-BDB9DB/2020\|EPI_ISL_709666\|20 | ···················· | ···················· | ···················· | ···················· | ······T············· | ···················· | |
| W | 4851 hCoV-19/England/MILK-BDC66A/2020\|EPI_ISL_709669\|20 | ···················· | ···················· | ···················· | ···················· | ···················· | ···················· | |
| W | 4852 hCoV-19/England/MILK-BDC776/2020\|EPI_ISL_709673\|20 | ···················· | ···················· | ···················· | ···················· | ···················· | ···················· | |
| W | 4853 hCoV-19/England/MILK-BDBD93/2020\|EPI_ISL_709674\|20 | ···················· | ···················· | ···················· | ···················· | ···················· | ···················· | |
| W | 4854 hCoV-19/England/MILK-BDC5A9/2020\|EPI_ISL_709675\|20 | ···················· | ···················· | ···················· | ···················· | ···················· | ···················· | |
| W | 4855 hCoV-19/England/MILK-BDC187/2020\|EPI_ISL_709676\|20 | ···················· | ···················· | ···················· | ···················· | ···················· | ···················· | |
| W | 4856 hCoV-19/England/MILK-BDC65B/2020\|EPI_ISL_709678\|20 | ···················· | ···················· | ···················· | ···················· | ···················· | ···················· | |
| W | 4857 hCoV-19/England/MILK-BDC327/2020\|EPI_ISL_709679\|20 | ···················· | ·A·················· | ···················· | ···················· | ···················· | ···················· | |
| W | 4858 hCoV-19/England/MILK-BDB56E/2020\|EPI_ISL_709680\|20 | ···················· | ···················· | ···················· | ···················· | ···················· | ···················· | |
| W | 4859 hCoV-19/England/MILK-BDBFBB/2020\|EPI_ISL_709684\|20 | ···················· | ···················· | ···················· | ···················· | ···················· | ···················· | |
| W | 4860 hCoV-19/England/MILK-BDB6D4/2020\|EPI_ISL_709687\|20 | ···················· | ···················· | ···················· | ···················· | ·················Y·· | ···················· | 1 |
| W | 4861 hCoV-19/England/MILK-BDCB01/2020\|EPI_ISL_709689\|20 | ···················· | ···················· | ···················· | ···················· | ···················· | ···················· | |
| W | 4862 hCoV-19/England/MILK-BDC73A/2020\|EPI_ISL_709692\|20 | ···················· | ···················· | ···················· | ···················· | ···················· | ···················· | |
| W | 4863 hCoV-19/England/MILK-BDC4AC/2020\|EPI_ISL_709696\|20 | ···················· | ···················· | ···················· | ···················· | ···················· | ···················· | |
| W | 4864 hCoV-19/England/MILK-BDC22A/2020\|EPI_ISL_709697\|20 | ···················· | ···················· | ···················· | ···················· | ···················· | ···················· | |
| W | 4865 hCoV-19/USA/AZ-TG305962/2020\|EPI_ISL_694301\|2020-0 | ···················· | ···················· | ···················· | ···················· | ···················· | ···················· | |
| W | 4866 hCoV-19/USA/AZ-TG305971/2020\|EPI_ISL_694303\|2020-0 | ···················· | ···················· | ···················· | ···················· | ···················· | ···················· | |
| W | 4867 hCoV-19/USA/AZ-TG305975/2020\|EPI_ISL_694304\|2020-0 | ···················· | ···················· | ···················· | ···················· | ···················· | ···················· | |
| W | 4868 hCoV-19/USA/AZ-TG305977/2020\|EPI_ISL_694305\|2020-0 | ···················· | ···················· | ···················· | ···················· | ···················· | ···················· | |
| W | 4869 hCoV-19/USA/AZ-TG305995/2020\|EPI_ISL_694307\|2020-0 | ···················· | ···················· | ···················· | ···················· | ···················· | ···················· | |
| W | 4870 hCoV-19/USA/AZ-TG305997/2020\|EPI_ISL_694308\|2020-0 | ···················· | ···················· | ···················· | ···················· | ···················· | ···················· | |
| W | 4871 hCoV-19/USA/AZ-TG305999/2020\|EPI_ISL_694309\|2020-0 | ···················· | ···················· | ···················· | ···················· | ···················· | ···················· | |
| W | 4872 hCoV-19/USA/AZ-TG306003/2020\|EPI_ISL_694311\|2020-0 | ···················· | ·············A······ | ···················· | ···················· | ···················· | ···················· | |
| W | 4873 hCoV-19/USA/AZ-TG306005/2020\|EPI_ISL_694312\|2020-0 | ···················· | ·············A······ | ···················· | ···················· | ···················· | ···················· | |
| W | 4874 hCoV-19/USA/AZ-TG306014/2020\|EPI_ISL_694314\|2020-0 | ···················· | ···················· | ···················· | ···················· | ···················· | ···················· | |
| W | 4875 hCoV-19/USA/AZ-TG306016/2020\|EPI_ISL_694315\|2020-0 | ···················· | ···················· | ···················· | ···················· | ···················· | ···················· | |
| W | 4876 hCoV-19/USA/AZ-TG308847/2020\|EPI_ISL_694316\|2020-0 | ···················· | ···················· | ···················· | ···················· | ···················· | ···················· | |
| W | 4877 hCoV-19/USA/AZ-TG310667/2020\|EPI_ISL_694317\|2020-0 | ···················· | ···················· | ···················· | ···················· | ···················· | ···················· | |
| W | 4878 hCoV-19/USA/AZ-TG312312/2020\|EPI_ISL_694320\|2020-0 | ···················· | ···················· | ···················· | ···················· | ···················· | ···················· | |
| W | 4879 hCoV-19/USA/AZ-TG312321/2020\|EPI_ISL_694321\|2020-0 | ···················· | ···················· | ···················· | ···················· | ···················· | ···················· | |
| W | 4880 hCoV-19/USA/AZ-TG312398/2020\|EPI_ISL_694322\|2020-0 | ···················· | ···················· | ···················· | ···················· | ···················· | ···················· | |
| W | 4881 hCoV-19/USA/AZ-TG312514/2020\|EPI_ISL_694323\|2020-0 | ···················· | ···················· | ···················· | ···················· | ···················· | ···················· | |
| W | 4882 hCoV-19/USA/AZ-TG312661/2020\|EPI_ISL_694324\|2020-0 | ···················· | ···················· | ···················· | ···················· | ···················· | ···················· | |
| W | 4883 hCoV-19/USA/AZ-TG312663/2020\|EPI_ISL_694325\|2020-0 | ···················· | ···················· | ···················· | ···················· | ···················· | ···················· | |
| W | 4884 hCoV-19/USA/AZ-TG314277/2020\|EPI_ISL_694328\|2020-0 | ···················· | ···················· | ···················· | ···················· | ···················· | ···················· | |
| W | 4885 hCoV-19/USA/AZ-TG333630/2020\|EPI_ISL_694359\|2020-0 | ···················· | ···················· | ···················· | ···················· | ···················· | ···················· | |
| W | 4886 hCoV-19/USA/AZ-TG333692/2020\|EPI_ISL_694360\|2020-0 | ···················· | ···················· | ···················· | ···················· | ···················· | ···················· | |
| W | 4887 hCoV-19/USA/AZ-TG333735/2020\|EPI_ISL_694361\|2020-0 | ···················· | ·A·················· | ···················· | ···················· | ···················· | ···················· | |
| W | 4888 hCoV-19/USA/AZ-TG333784/2020\|EPI_ISL_694362\|2020-0 | ···················· | ····T··············· | ···················· | ···················· | ···················· | ···················· | |
| W | 4889 hCoV-19/USA/AZ-TG335239/2020\|EPI_ISL_694366\|2020-0 | ···················· | ···················· | ···················· | ···················· | ···················· | ···················· | |
| W | 4890 hCoV-19/USA/AZ-TG337589/2020\|EPI_ISL_694370\|2020-0 | ···················· | ···················· | ···················· | ···················· | ···················· | ···················· | |
| W | 4891 hCoV-19/USA/AZ-TG337590/2020\|EPI_ISL_694371\|2020-0 | ···················· | ···················· | ···················· | ···················· | ···················· | ···················· | |
| W | 4892 hCoV-19/USA/AZ-TG341879/2020\|EPI_ISL_694379\|2020-0 | ···················· | ·A·················· | ···················· | ···················· | ···················· | ···················· | |
| W | 4893 hCoV-19/USA/AZ-TG342042/2020\|EPI_ISL_694381\|2020-0 | ···················· | ···················· | ···················· | ···················· | ···················· | ···················· | |
| W | 4894 hCoV-19/USA/AZ-TG342744/2020\|EPI_ISL_694382\|2020-0 | ···················· | ···················· | ···················· | ···················· | ···················· | ···················· | |
| W | 4895 hCoV-19/USA/AZ-TG343840/2020\|EPI_ISL_694383\|2020-0 | ···················· | ···················· | ···················· | ···················· | ···················· | ···················· | |
| W | 4896 hCoV-19/USA/AZ-TG344972/2020\|EPI_ISL_694386\|2020-0 | ···················· | ···················· | ···················· | ···················· | ···················· | ···················· | |
| W | 4897 hCoV-19/USA/AZ-TG345029/2020\|EPI_ISL_694387\|2020-0 | ···················· | ···················· | ···················· | ···················· | ···················· | ···················· | |
| W | 4898 hCoV-19/USA/AZ-TG345476/2020\|EPI_ISL_694389\|2020-0 | ···················· | ···················· | ···················· | ···················· | ···················· | ···················· | |
| W | 4899 hCoV-19/USA/AZ-TG345747/2020\|EPI_ISL_694391\|2020-0 | ···················· | ···················· | ···················· | ···················· | ···················· | ···················· | |
| W | 4900 hCoV-19/USA/AZ-TG345780/2020\|EPI_ISL_694392\|2020-0 | ···················· | ····T··············· | ···················· | ···················· | ···················· | ···················· | |
| W | 4901 hCoV-19/USA/AZ-TG347260/2020\|EPI_ISL_694394\|2020-0 | ···················· | ···················· | ···················· | ···················· | ···················· | ···················· | |
| W | 4902 hCoV-19/USA/AZ-TG348980/2020\|EPI_ISL_694398\|2020-0 | ···················· | ···················· | ···················· | ···················· | ···················· | ···················· | |
| W | 4903 hCoV-19/USA/AZ-TG349005/2020\|EPI_ISL_694399\|2020-0 | ···················· | ···················· | ···················· | ···················· | ···················· | ···················· | |
| W | 4904 hCoV-19/USA/AZ-TG569520/2020\|EPI_ISL_695600\|2020-0 | ···················· | ·A·················· | ···················· | ···················· | ···················· | ···················· | |
| W | 4905 hCoV-19/USA/AZ-TG569532/2020\|EPI_ISL_695608\|2020-0 | ···················· | ··C················· | ···················· | ···················· | ···················· | ···················· | |
| W | 4906 hCoV-19/USA/AZ-TG569537/2020\|EPI_ISL_695611\|2020-0 | ···················· | ···················· | ···················· | ···················· | ···················· | ···················· | |
| W | 4907 hCoV-19/USA/AZ-TG569539/2020\|EPI_ISL_695612\|2020-0 | ···················· | ··C················· | ···················· | ···················· | ···················· | ···················· | |
| W | 4908 hCoV-19/USA/AZ-TG572095/2020\|EPI_ISL_695621\|2020-0 | ···················· | ···················· | ···················· | ···················· | ···················· | ···················· | |
| W | 4909 hCoV-19/USA/AZ-TG573651/2020\|EPI_ISL_695623\|2020-0 | ···················· | ·A·················· | ···················· | ···················· | ···················· | ···················· | |
| W | 4910 hCoV-19/USA/AZ-TG577617/2020\|EPI_ISL_695624\|2020-0 | ···················· | ···················· | ···················· | ···················· | ···················· | ···················· | |
| W | 4911 hCoV-19/USA/AZ-TG588256/2020\|EPI_ISL_695634\|2020-0 | ···················· | ·A·················· | ···················· | ···················· | ···················· | ···················· | |
| W | 4912 hCoV-19/USA/AZ-TG618044/2020\|EPI_ISL_695686\|2020-0 | ···················· | ···················· | ···················· | ···················· | ···················· | ···················· | |
| W | 4913 hCoV-19/USA/AZ-TG618047/2020\|EPI_ISL_695689\|2020-0 | ···················· | ···················· | ···················· | ···················· | ···················· | ···················· | |
| W | 4914 hCoV-19/USA/AZ-TG618052/2020\|EPI_ISL_695693\|2020-0 | ···················· | ···················· | ···················· | ···················· | ···················· | ···················· | |
| W | 4915 hCoV-19/USA/AZ-TG618056/2020\|EPI_ISL_695696\|2020-0 | ···················· | ·A·················· | ···················· | ···················· | ···················· | ···················· | |
| W | 4916 hCoV-19/England/CAMC-BD1B48/2020\|EPI_ISL_702209\|20 | ···················· | ···················· | ···················· | ···················· | ···················· | ···················· | |
| W | 4917 hCoV-19/England/CAMC-BD2075/2020\|EPI_ISL_702210\|20 | ···················· | ···················· | ···················· | ···················· | ···················· | ···················· | |
| W | 4918 hCoV-19/England/CAMC-BD1AA5/2020\|EPI_ISL_702211\|20 | ···················· | ···················· | ···················· | ···················· | ···················· | ···················· | |
| W | 4919 hCoV-19/England/CAMC-BD227F/2020\|EPI_ISL_702212\|20 | ···················· | ···················· | ···················· | ···················· | ···················· | ···················· | |
| W | 4920 hCoV-19/England/CAMC-BD3690/2020\|EPI_ISL_702213\|20 | ···················· | ···················· | ···················· | ···················· | ···················· | ···················· | |
| W | 4921 hCoV-19/England/CAMC-BD20A2/2020\|EPI_ISL_702214\|20 | ···················· | ···················· | ···················· | ···················· | ···················· | ···················· | |
| W | 4922 hCoV-19/England/CAMC-BD16A1/2020\|EPI_ISL_702215\|20 | ···················· | ···················· | ···················· | ···················· | ···················· | ···················· | |
| W | 4923 hCoV-19/England/CAMC-BD4653/2020\|EPI_ISL_702216\|20 | ···················· | ···················· | ···················· | ···················· | ···················· | ···················· | |
| W | 4924 hCoV-19/England/CAMC-BD1E8B/2020\|EPI_ISL_702217\|20 | ···················· | ···················· | ···················· | ···················· | ···················· | ···················· | |
| W | 4925 hCoV-19/England/CAMC-BD21CD/2020\|EPI_ISL_702218\|20 | ···················· | ···················· | ···················· | ···················· | ···················· | ···················· | |
| W | 4926 hCoV-19/England/CAMC-BD1C72/2020\|EPI_ISL_702223\|20 | ···················· | ···················· | ···················· | ···················· | ···················· | ···················· | |
| W | 4927 hCoV-19/England/CAMC-BD1692/2020\|EPI_ISL_702224\|20 | ···················· | ···················· | ···················· | ···················· | ···················· | ···················· | |

| | | | | | | | | |
|---|---|---|---|---|---|---|---|---|
| W | 4928 | hCoV-19/England/CAMC-BD1AB4/2020\|EPI_ISL_702225\|20 | ···················· | ···················· | ····················· | ···················· | ······T············· | ···················· |
| W | 4929 | hCoV-19/England/CAMC-BD1E21/2020\|EPI_ISL_702227\|20 | ···················· | ···················· | ····················· | ···················· | ···················· | ···················· |
| W | 4930 | hCoV-19/England/CAMC-BD1E4F/2020\|EPI_ISL_702234\|20 | ···················· | ···················· | ····················· | ···················· | ···················· | ···················· |
| W | 4931 | hCoV-19/England/CAMC-BD2AB3/2020\|EPI_ISL_702235\|20 | ···················· | ···················· | ····················· | ···················· | ···················· | ···················· |
| W | 4932 | hCoV-19/England/CAMC-BD1B0C/2020\|EPI_ISL_702236\|20 | ···················· | ···················· | ····················· | ···················· | ···················· | ···················· |
| W | 4933 | hCoV-19/England/CAMC-BD186F/2020\|EPI_ISL_702240\|20 | ···················· | ···················· | ····················· | ···················· | ···················· | ···················· |
| W | 4934 | hCoV-19/England/CAMC-BD21DC/2020\|EPI_ISL_702241\|20 | ···················· | ···················· | ····················· | ···················· | ···················· | ···················· |
| W | 4935 | hCoV-19/England/CAMC-BD1AF0/2020\|EPI_ISL_702242\|20 | ···················· | ···················· | ····················· | ···················· | ···················· | ···················· |
| W | 4936 | hCoV-19/England/CAMC-BD4671/2020\|EPI_ISL_702245\|20 | ···················· | ···················· | ····················· | ···················· | ···················· | ···················· |
| W | 4937 | hCoV-19/England/CAMC-BD3663/2020\|EPI_ISL_702251\|20 | ···················· | ·A·················· | ····················· | ···················· | ···················· | ···················· |
| W | 4938 | hCoV-19/England/CAMC-BD45CF/2020\|EPI_ISL_702252\|20 | ···················· | ···················· | ····················· | ···················· | ···················· | ···················· |
| W | 4939 | hCoV-19/England/CAMC-BD1F5B/2020\|EPI_ISL_702257\|20 | ···················· | ···················· | ····················· | ···················· | ···················· | ···················· |
| W | 4940 | hCoV-19/England/CAMC-BD43C5/2020\|EPI_ISL_702259\|20 | ···················· | ···················· | ····················· | ···················· | ···················· | ···················· |
| W | 4941 | hCoV-19/England/CAMC-BD17CC/2020\|EPI_ISL_702261\|20 | ···················· | ···················· | ····················· | ···················· | ···················· | ···················· |
| W | 4942 | hCoV-19/England/CAMC-BD3A2B/2020\|EPI_ISL_702262\|20 | ···················· | ···················· | ····················· | ···················· | ···················· | ···················· |
| W | 4943 | hCoV-19/England/CAMC-BD1771/2020\|EPI_ISL_702264\|20 | ···················· | ···················· | ····················· | ···················· | ···················· | ···················· |
| W | 4944 | hCoV-19/England/CAMC-BD1823/2020\|EPI_ISL_702266\|20 | ···················· | ···················· | ····················· | ···················· | ···················· | ···················· |
| W | 4945 | hCoV-19/England/CAMC-BD20EE/2020\|EPI_ISL_702267\|20 | ···················· | ···················· | ····················· | ···················· | ···················· | ···················· |
| W | 4946 | hCoV-19/England/CAMC-BD48E4/2020\|EPI_ISL_702269\|20 | ···················· | ···················· | ····················· | ···················· | ···················· | ···················· |
| W | 4947 | hCoV-19/England/CAMC-BD36AF/2020\|EPI_ISL_702271\|20 | ···················· | ···················· | ····················· | ···················· | ···················· | ···················· |
| W | 4948 | hCoV-19/England/CAMC-BD2AA4/2020\|EPI_ISL_702272\|20 | ···················· | ···················· | ····················· | ···················· | ···················· | ···················· |
| W | 4949 | hCoV-19/England/CAMC-BD14F2/2020\|EPI_ISL_702273\|20 | ···················· | ···················· | ····················· | ···················· | ···················· | ···················· |
| W | 4950 | hCoV-19/England/CAMC-BD28D7/2020\|EPI_ISL_702278\|20 | ···················· | ···················· | ····················· | ···················· | ···················· | ···················· |
| W | 4951 | hCoV-19/England/CAMC-BD2084/2020\|EPI_ISL_702279\|20 | ···················· | ···················· | ····················· | ···················· | ···················· | ···················· |
| W | 4952 | hCoV-19/England/CAMC-BD154A/2020\|EPI_ISL_702280\|20 | ···················· | ·A·················· | ····················· | ···················· | ···················· | ···················· |
| W | 4953 | hCoV-19/England/CAMC-BD2109/2020\|EPI_ISL_702285\|20 | ···················· | ···················· | ····················· | ···················· | ···················· | ···················· |
| W | 4954 | hCoV-19/England/CAMC-BD48A8/2020\|EPI_ISL_702286\|20 | ···················· | ···················· | ····················· | ···················· | ···················· | ···················· |
| W | 4955 | hCoV-19/England/CAMC-BD1C09/2020\|EPI_ISL_702287\|20 | ···················· | ···················· | ····················· | ···················· | ···················· | ···················· |
| W | 4956 | hCoV-19/England/CAMC-BD1E9A/2020\|EPI_ISL_702288\|20 | ···················· | ···················· | ····················· | ···················· | ···················· | ···················· |
| W | 4957 | hCoV-19/England/CAMC-BD444A/2020\|EPI_ISL_702289\|20 | ···················· | ···················· | ····················· | ···················· | ···················· | ···················· |
| W | 4958 | hCoV-19/England/CAMC-BD16ED/2020\|EPI_ISL_702292\|20 | ···················· | ·A·················· | ····················· | ···················· | ···················· | ··A················· |
| W | 4959 | hCoV-19/England/CAMC-BD2AD1/2020\|EPI_ISL_702293\|20 | ···················· | ···················· | ····················· | ···················· | ······T············· | ···················· |
| W | 4960 | hCoV-19/England/CAMC-BD48C6/2020\|EPI_ISL_702294\|20 | ···················· | ···················· | ····················· | ···················· | ···················· | ···················· |
| W | 4961 | hCoV-19/England/CAMC-BD187E/2020\|EPI_ISL_702295\|20 | ···················· | ···················· | ····················· | ···················· | ···················· | ···················· |
| W | 4962 | hCoV-19/England/CAMC-BD1EA9/2020\|EPI_ISL_702296\|20 | ···················· | ···················· | ····················· | ···················· | ······T············· | ···················· |
| W | 4963 | hCoV-19/England/CAMC-BD20B1/2020\|EPI_ISL_702298\|20 | ···················· | ·A·················· | ····················· | ···················· | ···················· | ··A················· |
| W | 4964 | hCoV-19/England/CAMC-BD14B6/2020\|EPI_ISL_702299\|20 | ···················· | ·A·················· | ····················· | ···················· | ···················· | ···················· |
| W | 4965 | hCoV-19/England/QEUH-BC428F/2020\|EPI_ISL_703514\|20 | ···················· | ···················· | ····················· | ···················· | ···················· | ···················· |
| W | 4966 | hCoV-19/England/QEUH-BC4762/2020\|EPI_ISL_703515\|20 | ···················· | ···················· | ····················· | ···················· | ···················· | ···················· |
| W | 4967 | hCoV-19/England/QEUH-BC4137/2020\|EPI_ISL_703517\|20 | ···················· | ···················· | ····················· | ···················· | ···················· | ···················· |
| W | 4968 | hCoV-19/England/QEUH-BC460B/2020\|EPI_ISL_703541\|20 | ···················· | ···················· | ····················· | ···················· | ···················· | ···················· |
| W | 4969 | hCoV-19/England/ALDP-BCA975/2020\|EPI_ISL_703543\|20 | ···················· | ·········T·········· | ····················· | ···················· | ···················· | ···················· |
| W | 4970 | hCoV-19/England/QEUH-BC3FD4/2020\|EPI_ISL_703546\|20 | ···················· | ···················· | ····················· | ···················· | ······T············· | ···················· |
| W | 4971 | hCoV-19/England/ALDP-BCA85A/2020\|EPI_ISL_703547\|20 | ···················· | ···················· | ····················· | ···················· | ···················· | ···················· |
| W | 4972 | hCoV-19/England/ALDP-BCAB42/2020\|EPI_ISL_703549\|20 | ···················· | ···················· | ····················· | ···················· | ···················· | ···················· |
| W | 4973 | hCoV-19/England/ALDP-BCA2F2/2020\|EPI_ISL_703552\|20 | ···················· | ···················· | ····················· | ···················· | ···················· | ···················· |
| W | 4974 | hCoV-19/England/QEUH-BC46FC/2020\|EPI_ISL_703553\|20 | ···················· | ···················· | ····················· | ···················· | ···················· | ···················· |
| W | 4975 | hCoV-19/England/ALDP-BCA298/2020\|EPI_ISL_703555\|20 | ···················· | ···················· | ····················· | ···················· | ···················· | ···················· |
| W | 4976 | hCoV-19/England/QEUH-BC46A1/2020\|EPI_ISL_703557\|20 | ···················· | ···················· | ····················· | ···················· | ···················· | ···················· |
| W | 4977 | hCoV-19/England/ALDP-BCAA09/2020\|EPI_ISL_703558\|20 | ···················· | ···················· | ····················· | ···················· | ···················· | ···················· |
| W | 4978 | hCoV-19/England/QEUH-BC3FE3/2020\|EPI_ISL_703560\|20 | ···················· | ···················· | ····················· | ···················· | ···················· | ···················· |
| W | 4979 | hCoV-19/England/QEUH-BC41DD/2020\|EPI_ISL_703561\|20 | ···················· | ···················· | ····················· | ···················· | ···················· | ···················· |
| W | 4980 | hCoV-19/England/ALDP-BCA19B/2020\|EPI_ISL_703563\|20 | ···················· | ···················· | ····················· | ···················· | ···················· | ···················· |
| W | 4981 | hCoV-19/England/ALDP-BCA81E/2020\|EPI_ISL_703565\|20 | ···················· | ···················· | ····················· | ···················· | ·······T············ | ···················· |
| W | 4982 | hCoV-19/England/ALDP-BCA623/2020\|EPI_ISL_703566\|20 | ···················· | ···················· | ····················· | ···················· | ···················· | ···················· |
| W | 4983 | hCoV-19/England/ALDP-BCA23E/2020\|EPI_ISL_703568\|20 | ···················· | ···················· | ····················· | ···················· | ···················· | ···················· |
| W | 4984 | hCoV-19/England/ALDP-BCA605/2020\|EPI_ISL_703571\|20 | ···················· | ···················· | ····················· | ···················· | ···················· | ···················· |
| W | 4985 | hCoV-19/England/ALDP-BCA508/2020\|EPI_ISL_703573\|20 | ···················· | ···················· | ····················· | ···················· | ···················· | ··A················· |
| W | 4986 | hCoV-19/England/ALDP-BCAA90/2020\|EPI_ISL_703576\|20 | ···················· | ···················· | ····················· | ···················· | ···················· | ···················· |
| W | 4987 | hCoV-19/England/QEUH-BC446B/2020\|EPI_ISL_703578\|20 | ···················· | ···················· | ····················· | ···················· | ···················· | ···················· |
| W | 4988 | hCoV-19/England/ALDP-BCA016/2020\|EPI_ISL_703580\|20 | ···················· | ···················· | ····················· | ···················· | ···················· | ···················· |
| W | 4989 | hCoV-19/England/ALDP-BCA8A5/2020\|EPI_ISL_703581\|20 | ···················· | ···················· | ····················· | ···················· | ···················· | ···················· |
| W | 4990 | hCoV-19/England/QEUH-BC4207/2020\|EPI_ISL_703583\|20 | ···················· | ···················· | ····················· | ···················· | ···················· | ···················· |
| W | 4991 | hCoV-19/England/QEUH-BC4146/2020\|EPI_ISL_703584\|20 | ···················· | ···················· | ····················· | ···················· | ···················· | ···················· |
| W | 4992 | hCoV-19/England/ALDP-BCA33B/2020\|EPI_ISL_703586\|20 | ···················· | ···················· | ····················· | ···················· | ···················· | ···················· |
| W | 4993 | hCoV-19/England/ALDP-BCA6BA/2020\|EPI_ISL_703588\|20 | ···················· | ···················· | ····················· | ···················· | ···················· | ···················· |
| W | 4994 | hCoV-19/England/ALDP-BCA59F/2020\|EPI_ISL_703591\|20 | ···················· | ···················· | ····················· | ···················· | ···················· | ···················· |
| W | 4995 | hCoV-19/England/ALDP-BCAA18/2020\|EPI_ISL_703592\|20 | ···················· | ···················· | ····················· | ···················· | ···················· | ···················· |
| W | 4996 | hCoV-19/England/ALDP-BCA966/2020\|EPI_ISL_703594\|20 | ···················· | ···················· | ····················· | ···················· | ···················· | ···················· |
| W | 4997 | hCoV-19/England/ALDP-BCA3A4/2020\|EPI_ISL_703595\|20 | ···················· | ···················· | ····················· | ···················· | ···················· | ···················· |
| W | 4998 | hCoV-19/England/ALDP-BCA359/2020\|EPI_ISL_703597\|20 | ···················· | ···················· | ····················· | ···················· | ···················· | ···················· |
| W | 4999 | hCoV-19/England/QEUH-BC48E7/2020\|EPI_ISL_703598\|20 | ···················· | ···················· | ····················· | ···················· | ···················· | ···················· |
| W | 5000 | hCoV-19/England/MILK-A91BBE/2020\|EPI_ISL_704845\|20 | ···················· | ···················· | ····················· | ···················· | ···················· | ···················· |
| W | 5001 | hCoV-19/England/CAMC-BC05A8/2020\|EPI_ISL_704846\|20 | ···················· | ···················· | ····················· | ···················· | ···················· | ···················· |
| W | 5003 | hCoV-19/England/CAMC-BBFFEA/2020\|EPI_ISL_704848\|20 | ···················· | ···················· | ····················· | ···················· | ······T············· | ···················· |
| W | 5004 | hCoV-19/England/MILK-B3A6F1/2020\|EPI_ISL_704872\|20 | ···················· | ·A·················· | ····················· | ···················· | ···················· | ··A················· |
| W | 5005 | hCoV-19/England/MILK-B3C6B3/2020\|EPI_ISL_704874\|20 | ···················· | ·A·················· | ····················· | ···················· | ···················· | ··A················· |
| W | 5006 | hCoV-19/England/MILK-B3CB69/2020\|EPI_ISL_704877\|20 | ···················· | ·A·················· | ····················· | ···················· | ···················· | ··A················· |
| W | 5007 | hCoV-19/England/MILK-B3CFA9/2020\|EPI_ISL_704879\|20 | ···················· | ·A·················· | ····················· | ···················· | ···················· | ··A················· |
| W | 5008 | hCoV-19/England/CAMC-BC05E4/2020\|EPI_ISL_704880\|20 | ···················· | ···················· | ····················· | ···················· | ···················· | ···················· |

| | | | | | | | |
|---|---|---|---|---|---|---|---|
| W | 5009 hCoV-19/England/CAMC-BC1BF0/2020\|EPI_ISL_704881\|20 | ···················· | ···················· | ···················· | ···················· | ···················· | ···················· |
| W | 5010 hCoV-19/England/MILK-B3D5C4/2020\|EPI_ISL_704882\|20 | ···················· | ·A·················· | ···················· | ···················· | ···················· | ··A················· |
| W | 5011 hCoV-19/England/CAMC-BC22FA/2020\|EPI_ISL_704883\|20 | ···················· | ···················· | ···················· | ···················· | ···················· | ···················· |
| W | 5012 hCoV-19/England/MILK-B3DB95/2020\|EPI_ISL_704885\|20 | ···················· | ·A·················· | ···················· | ···················· | ···················· | ··A················· |
| W | 5013 hCoV-19/England/CAMC-BC07B1/2020\|EPI_ISL_704886\|20 | ···················· | ···················· | ···················· | ···················· | ···················· | ···················· |
| W | 5014 hCoV-19/England/NORW-EEE2C/2020\|EPI_ISL_704887\|202 | ···················· | ·A·················· | ···················· | ···················· | ···················· | ··A················· |
| W | 5015 hCoV-19/England/CAMC-BC0195/2020\|EPI_ISL_704888\|20 | ···················· | ···················· | ···················· | ···················· | ···················· | ···················· |
| W | 5016 hCoV-19/England/CAMC-BC0BC4/2020\|EPI_ISL_704889\|20 | ···················· | ···················· | ···················· | ···················· | ···················· | ···················· |
| W | 5017 hCoV-19/England/NORW-EEE3B/2020\|EPI_ISL_704890\|202 | ···················· | ·A·················· | ···················· | ···················· | ···················· | ··A················· |
| W | 5018 hCoV-19/England/CAMC-BC02CF/2020\|EPI_ISL_704891\|20 | ···················· | ···················· | ···················· | ···················· | ···················· | ···················· |
| W | 5019 hCoV-19/England/NORW-F642F/2020\|EPI_ISL_704892\|202 | ···················· | ·A·················· | ···················· | ···················· | ···················· | ··A················· |
| W | 5020 hCoV-19/England/CAMC-BC07C0/2020\|EPI_ISL_704893\|20 | ···················· | ···················· | ···················· | ···················· | ···················· | ···················· |
| W | 5021 hCoV-19/England/CAMC-BC0E16/2020\|EPI_ISL_704894\|20 | ···················· | ···················· | ···················· | ···················· | ···················· | ···················· |
| W | 5022 hCoV-19/England/CAMC-BC06F0/2020\|EPI_ISL_704896\|20 | ···················· | ···················· | ···················· | ···················· | ···················· | ···················· |
| W | 5023 hCoV-19/USA/DE-DHSS-F994288/2020\|EPI_ISL_708433\|20 | ···················· | ···················· | ···················· | ···················· | ···················· | ···················· |
| W | 5024 hCoV-19/USA/DE-DHSS-F1001769/2020\|EPI_ISL_708434\|2 | ···················· | ···················· | ···················· | ···················· | ···················· | ···················· |
| W | 5025 hCoV-19/USA/DE-DHSS-F994223/2020\|EPI_ISL_708435\|20 | ···················· | ···················· | ···················· | ···················· | ···················· | ···················· |
| W | 5026 hCoV-19/USA/DE-DHSS-F995530/2020\|EPI_ISL_708436\|20 | ···················· | ···········C········ | ···················· | ···················· | ···················· | ···················· |
| W | 5027 hCoV-19/USA/DE-DHSS-F1019096/2020\|EPI_ISL_708437\|2 | ···················· | ···················· | ···················· | ···················· | ···················· | ···················· |
| W | 5028 hCoV-19/USA/DE-DHSS-F1019125/2020\|EPI_ISL_708438\|2 | ···················· | ·A·················· | ···················· | ···················· | ···················· | ···················· |
| W | 5029 hCoV-19/USA/DE-DHSS-F1018204/2020\|EPI_ISL_708439\|2 | ···················· | ···················· | ···················· | ···················· | ···················· | ···················· |
| W | 5030 hCoV-19/USA/DE-DHSS-F986368/2020\|EPI_ISL_708440\|20 | ···················· | ···················· | ···················· | ···················· | ···················· | ···················· |
| W | 5031 hCoV-19/USA/DE-DHSS-F970980/2020\|EPI_ISL_708441\|20 | ···················· | ···················· | ···················· | ···················· | ···················· | ···················· |
| W | 5032 hCoV-19/USA/DE-DHSS-F1019139/2020\|EPI_ISL_708442\|2 | ···················· | ···················· | ···················· | ···················· | ···················· | ···················· |
| W | 5033 hCoV-19/USA/DE-DHSS-F1020819/2020\|EPI_ISL_708443\|2 | ···················· | ···················· | ···················· | ···················· | ···················· | ···················· |
| W | 5034 hCoV-19/USA/DE-DHSS-F1022209/2020\|EPI_ISL_708444\|2 | ···················· | ···················· | ···················· | ···················· | ···················· | ···················· |
| W | 5035 hCoV-19/USA/DE-DHSS-F956504/2020\|EPI_ISL_708445\|20 | ···················· | ···················· | ···················· | ···················· | ···················· | ···················· |
| W | 5036 hCoV-19/USA/DE-DHSS-F995648/2020\|EPI_ISL_708446\|20 | ···················· | ·A·················· | ···················· | ···················· | ···················· | ···················· |
| W | 5037 hCoV-19/USA/DE-DHSS-F1019092/2020\|EPI_ISL_708447\|2 | ···················· | ···················· | ···················· | ···················· | ···················· | ···················· |
| W | 5038 hCoV-19/USA/DE-DHSS-F980816/2020\|EPI_ISL_708448\|20 | ···················· | ···········C········ | ···················· | ···················· | ···················· | ···················· |
| W | 5039 hCoV-19/USA/DE-DHSS-F1017497/2020\|EPI_ISL_708449\|2 | ···················· | ·A·················· | ···················· | ···················· | ···················· | ···················· |
| W | 5040 hCoV-19/USA/DE-DHSS-F1020844/2020\|EPI_ISL_708450\|2 | ···················· | ···················· | ···················· | ···················· | ···················· | ···················· |
| W | 5041 hCoV-19/USA/DE-DHSS-F1020823/2020\|EPI_ISL_708451\|2 | ···················· | ···················· | ···················· | ···················· | ···················· | ···················· |
| W | 5042 hCoV-19/USA/DE-DHSS-F1020918/2020\|EPI_ISL_708452\|2 | ···················· | ·A·················· | ···················· | ···················· | ···················· | ···················· |
| W | 5043 hCoV-19/Belgium/ULG-10967/2020\|EPI_ISL_708453\|2020 | ···················· | ···················· | ···················· | ···················· | ···················· | ···················· |
| W | 5044 hCoV-19/Belgium/ULG-10969/2020\|EPI_ISL_708454\|2020 | ···················· | ···················· | ···················· | ···················· | ···················· | ···················· |
| W | 5045 hCoV-19/Belgium/ULG-10986/2020\|EPI_ISL_708455\|2020 | ···················· | ···················· | ···················· | ···················· | ···················· | ···················· |
| W | 5046 hCoV-19/Belgium/ULG-11014/2020\|EPI_ISL_708458\|2020 | ···················· | ···················· | ···················· | ···················· | ···················· | ···················· |
| W | 5047 hCoV-19/Belgium/ULG-11015/2020\|EPI_ISL_708459\|2020 | ···················· | ···················· | ···················· | ···················· | ···················· | ···················· |
| W | 5048 hCoV-19/USA/MN-MDH-2122/2020\|EPI_ISL_708460\|2020-1 | ···················· | ···················· | ···················· | ···················· | ···················· | ···················· |
| W | 5049 hCoV-19/USA/MN-MDH-2123/2020\|EPI_ISL_708461\|2020-1 | ···················· | ···················· | ···················· | ···················· | ···················· | ···················· |
| W | 5050 hCoV-19/USA/MN-MDH-2124/2020\|EPI_ISL_708462\|2020-1 | ········T··········· | ·A·················· | ···················· | ···················· | ···················· | ···················· |
| W | 5051 hCoV-19/USA/MN-MDH-2125/2020\|EPI_ISL_708463\|2020-1 | ···················· | ···················· | ···················· | ···················· | ···················· | ···················· |
| W | 5052 hCoV-19/USA/MN-MDH-2126/2020\|EPI_ISL_708464\|2020-1 | ···················· | ···················· | ···················· | ···················· | ···················· | ···················· |
| W | 5053 hCoV-19/USA/MN-MDH-2127/2020\|EPI_ISL_708465\|2020-1 | ···················· | ···················· | ···················· | ···················· | ···················· | ···················· |
| W | 5054 hCoV-19/USA/MN-MDH-2128/2020\|EPI_ISL_708466\|2020-1 | ···················· | ···················· | ···················· | ···················· | ···················· | ···················· |
| W | 5055 hCoV-19/USA/MN-MDH-2129/2020\|EPI_ISL_708467\|2020-1 | ···················· | ···················· | ···················· | ···················· | ···················· | ···················· |
| W | 5056 hCoV-19/USA/MN-MDH-2130/2020\|EPI_ISL_708468\|2020-1 | ···················· | ···················· | ···················· | ···················· | ···················· | ···················· |
| W | 5057 hCoV-19/USA/MN-MDH-2131/2020\|EPI_ISL_708469\|2020-1 | ···················· | ···················· | ···················· | ···················· | ···················· | ···················· |
| W | 5058 hCoV-19/USA/MN-MDH-2152/2020\|EPI_ISL_708490\|2020-1 | ···················· | ···················· | ···················· | ···················· | ···················· | ···················· |
| W | 5059 hCoV-19/USA/MN-MDH-2153/2020\|EPI_ISL_708491\|2020-1 | ···················· | ··C················· | ···················· | ···················· | ···················· | ···················· |
| W | 5060 hCoV-19/USA/MN-MDH-2154/2020\|EPI_ISL_708492\|2020-1 | ···················· | ···················· | ···················· | ···················· | ···················· | ···················· |
| W | 5061 hCoV-19/USA/MN-MDH-2155/2020\|EPI_ISL_708493\|2020-1 | ···················· | ···················· | ···················· | ···················· | ···················· | ···················· |
| W | 5062 hCoV-19/USA/MN-MDH-2156/2020\|EPI_ISL_708494\|2020-1 | ···················· | ···················· | ···················· | ···················· | ···················· | ···················· |
| W | 5063 hCoV-19/USA/MN-MDH-2157/2020\|EPI_ISL_708495\|2020-1 | ···················· | ···················· | ···················· | ···················· | ···················· | ···················· |
| W | 5064 hCoV-19/USA/MN-MDH-2158/2020\|EPI_ISL_708496\|2020-1 | ···················· | ···················· | ···················· | ···················· | ···················· | ···················· |
| W | 5065 hCoV-19/USA/MN-MDH-2159/2020\|EPI_ISL_708497\|2020-1 | ···················· | ···················· | ···················· | ···················· | ···················· | ···················· |
| W | 5066 hCoV-19/USA/MN-MDH-2160/2020\|EPI_ISL_708498\|2020-1 | ···················· | ···················· | ···················· | ···················· | ···················· | ···················· |
| W | 5067 hCoV-19/USA/MN-MDH-2161/2020\|EPI_ISL_708499\|2020-1 | ···················· | ···················· | ···················· | ···················· | ···················· | ···················· |
| W | 5068 hCoV-19/England/MILK-BDB7D1/2020\|EPI_ISL_709700\|20 | ···················· | ···················· | ···················· | ···················· | ···················· | ···················· |
| W | 5069 hCoV-19/England/MILK-BDBB8A/2020\|EPI_ISL_709701\|20 | ···················· | ···················· | ···················· | ···················· | ···················· | ···················· |
| W | 5070 hCoV-19/England/MILK-BDC20C/2020\|EPI_ISL_709703\|20 | ···················· | ···················· | ···················· | ···················· | ···················· | ···················· |
| W | 5071 hCoV-19/England/MILK-BDCAD7/2020\|EPI_ISL_709704\|20 | ···················· | ···················· | ···················· | ···················· | ···················· | ···················· |
| W | 5072 hCoV-19/England/MILK-BDB8B0/2020\|EPI_ISL_709705\|20 | ···················· | ·A·················· | ···················· | ···················· | ···················· | ···················· |
| W | 5073 hCoV-19/England/MILK-BDCA40/2020\|EPI_ISL_709710\|20 | ···················· | ···················· | ···················· | ···················· | ···················· | ···················· |
| W | 5074 hCoV-19/England/MILK-BDC749/2020\|EPI_ISL_709711\|20 | ···················· | ···················· | ···················· | ···················· | ···················· | ···················· |
| W | 5075 hCoV-19/England/MILK-BDC3AF/2020\|EPI_ISL_709713\|20 | ···················· | ···················· | ···················· | ···················· | ···················· | ···················· |
| W | 5076 hCoV-19/England/MILK-BDB74A/2020\|EPI_ISL_709716\|20 | ···················· | ···················· | ···················· | ···················· | ···················· | ···················· |
| W | 5077 hCoV-19/England/MILK-BDC512/2020\|EPI_ISL_709719\|20 | ···················· | ···················· | ···················· | ···················· | ···················· | ···················· |
| W | 5078 hCoV-19/England/MILK-BDC785/2020\|EPI_ISL_709721\|20 | ···················· | ···················· | ···················· | ···················· | ···················· | ···················· |
| W | 5079 hCoV-19/England/MILK-BDC406/2020\|EPI_ISL_709723\|20 | ···················· | ···················· | ···················· | ···················· | ···················· | ··A················· |
| W | 5080 hCoV-19/England/MILK-BDC49D/2020\|EPI_ISL_709725\|20 | ···················· | ·A·················· | ···················· | ···················· | ···················· | ···················· |
| W | 5081 hCoV-19/England/MILK-BDC248/2020\|EPI_ISL_709727\|20 | ···················· | ···················· | ···················· | ···················· | ···················· | ···················· |
| W | 5082 hCoV-19/England/MILK-BDB917/2020\|EPI_ISL_709731\|20 | ···················· | ···················· | ···················· | ···················· | ···················· | ···················· |
| W | 5083 hCoV-19/England/MILK-BDB73B/2020\|EPI_ISL_709733\|20 | ···················· | ···················· | ···················· | ···················· | ···················· | ···················· |
| W | 5084 hCoV-19/England/MILK-BDC2C0/2020\|EPI_ISL_709734\|20 | ···················· | ···················· | ···················· | ···················· | ·····T·············· | ···················· |
| W | 5085 hCoV-19/England/MILK-BDB6A7/2020\|EPI_ISL_709735\|20 | ···················· | ···················· | ···················· | ···················· | ···················· | ···················· |
| W | 5086 hCoV-19/England/MILK-BDB698/2020\|EPI_ISL_709738\|20 | ···················· | ···················· | ···················· | ···················· | ···················· | ···················· |
| W | 5087 hCoV-19/England/MILK-BDC62E/2020\|EPI_ISL_709740\|20 | ···················· | ···················· | ···················· | ···················· | ···················· | ···················· |
| W | 5088 hCoV-19/England/MILK-BDBCA5/2020\|EPI_ISL_709741\|20 | ···················· | ·A·················· | ···················· | ···················· | ···················· | ··A················· |

| | | | | | | | |
|---|---|---|---|---|---|---|---|
| W | 5089 hCoV-19/England/MILK-BDBDB1/2020\|EPI_ISL_709742\|20 | ···················· | ···················· | ····················· | ···················· | ···················· | ···················· |
| W | 5090 hCoV-19/England/MILK-BDB892/2020\|EPI_ISL_709743\|20 | ···················· | ···················· | ····················· | ···················· | ···················· | ···················· |
| W | 5091 hCoV-19/England/MILK-BDC71C/2020\|EPI_ISL_709747\|20 | ···················· | ···················· | ····················· | ···················· | ···················· | ···················· |
| W | 5092 hCoV-19/England/MILK-BDB908/2020\|EPI_ISL_709749\|20 | ···················· | ···················· | ····················· | ···················· | ···················· | ···················· |
| W | 5093 hCoV-19/England/MILK-BDC758/2020\|EPI_ISL_709750\|20 | ···················· | ···················· | ····················· | ···················· | ···················· | ···················· |
| W | 5094 hCoV-19/England/MILK-BDC9F8/2020\|EPI_ISL_709751\|20 | ···················· | ···················· | ····················· | ···················· | ···················· | ···················· |
| W | 5095 hCoV-19/England/MILK-BDBA05/2020\|EPI_ISL_709754\|20 | ···················· | ···················· | ····················· | ···················· | ···················· | ···················· |
| W | 5096 hCoV-19/England/MILK-BDB7B3/2020\|EPI_ISL_709757\|20 | ···················· | ···················· | ····················· | ···················· | ···················· | ···················· |
| W | 5097 hCoV-19/England/MILK-BDC13C/2020\|EPI_ISL_709760\|20 | ···················· | ···················· | ····················· | ···················· | ·····T·············· | ···················· |
| W | 5098 hCoV-19/England/MILK-BDBF15/2020\|EPI_ISL_709761\|20 | ···················· | ···················· | ····················· | ···················· | ···················· | ···················· |
| W | 5099 hCoV-19/England/MILK-BDCB10/2020\|EPI_ISL_709762\|20 | ···················· | ·A·················· | ····················· | ···················· | ···················· | ··A················· |
| W | 5100 hCoV-19/England/MILK-BDB65C/2020\|EPI_ISL_709764\|20 | ···················· | ···················· | ····················· | ···················· | ···················· | ···················· |
| W | 5101 hCoV-19/England/MILK-BDC64C/2020\|EPI_ISL_709766\|20 | ···················· | ···················· | ····················· | ···················· | ·····T·············· | ···················· |
| W | 5102 hCoV-19/England/MILK-BDB6F2/2020\|EPI_ISL_709768\|20 | ···················· | ···················· | ····················· | ···················· | ···················· | ···················· |
| W | 5103 hCoV-19/England/MILK-BDCAB9/2020\|EPI_ISL_709769\|20 | ···················· | ···················· | ····················· | ···················· | ···················· | ···················· |
| W | 5104 hCoV-19/England/MILK-BDC5E5/2020\|EPI_ISL_709770\|20 | ···················· | ···················· | ····················· | ···················· | ···················· | ···················· |
| W | 5105 hCoV-19/England/MILK-BDB59B/2020\|EPI_ISL_709771\|20 | ···················· | ···················· | ····················· | ···················· | ·····T·············· | ···················· |
| W | 5106 hCoV-19/England/MILK-BDB70E/2020\|EPI_ISL_709772\|20 | ···················· | ···················· | ····················· | ···················· | ···················· | ···················· |
| W | 5107 hCoV-19/England/MILK-BDB786/2020\|EPI_ISL_709774\|20 | ···················· | ···················· | ····················· | ···················· | ···················· | ···················· |
| W | 5108 hCoV-19/England/MILK-BDCAC8/2020\|EPI_ISL_709777\|20 | ···················· | ···················· | ····················· | ···················· | ···················· | ···················· |
| W | 5109 hCoV-19/England/MILK-BDBF06/2020\|EPI_ISL_709780\|20 | ···················· | ···················· | ····················· | ···················· | ·····T·············· | ···················· |
| W | 5110 hCoV-19/England/MILK-BDCAF5/2020\|EPI_ISL_709781\|20 | ···················· | ···················· | ····················· | ···················· | ·····T·············· | ···················· |
| W | 5111 hCoV-19/England/MILK-BDC6C4/2020\|EPI_ISL_709784\|20 | ···················· | ···················· | ····················· | ···················· | ···················· | ···················· |
| W | 5112 hCoV-19/England/MILK-BDC7C1/2020\|EPI_ISL_709785\|20 | ···················· | ···················· | ····················· | ···················· | ···················· | ···················· |
| W | 5113 hCoV-19/England/MILK-BDB71D/2020\|EPI_ISL_709786\|20 | ···················· | ···················· | ····················· | ···················· | ···················· | ···················· |
| W | 5114 hCoV-19/England/MILK-BDCA9B/2020\|EPI_ISL_709788\|20 | ···················· | ···················· | ····················· | ···················· | ···················· | ···················· |
| W | 5115 hCoV-19/England/MILK-BDC600/2020\|EPI_ISL_709789\|20 | ···················· | ···················· | ····················· | ···················· | ···················· | ···················· |
| W | 5116 hCoV-19/England/MILK-BDC688/2020\|EPI_ISL_709790\|20 | ···················· | ···················· | ····················· | ···················· | ···················· | ···················· |
| W | 5117 hCoV-19/England/MILK-BDC239/2020\|EPI_ISL_709792\|20 | ···················· | ···················· | ····················· | ···················· | ···················· | ···················· |
| W | 5118 hCoV-19/England/MILK-BDBFF7/2020\|EPI_ISL_709794\|20 | ···················· | ···················· | ····················· | ···················· | ···················· | ···················· |
| W | 5119 hCoV-19/England/MILK-BDC21B/2020\|EPI_ISL_709795\|20 | ···················· | ·A·················· | ····················· | ···················· | ···················· | ··A················· |
| W | 5120 hCoV-19/England/MILK-BDC6D3/2020\|EPI_ISL_709798\|20 | ···················· | ···················· | ····················· | ···················· | ···················· | ···················· |
| W | 5121 hCoV-19/USA/AZ-TG320856/2020\|EPI_ISL_694332\|2020-0 | ···················· | ···················· | ····················· | ···················· | ···················· | ···················· |
| W | 5122 hCoV-19/USA/AZ-TG320868/2020\|EPI_ISL_694333\|2020-0 | ···················· | ···················· | ····················· | ···················· | ···················· | ···················· |
| W | 5123 hCoV-19/USA/AZ-TG321300/2020\|EPI_ISL_694335\|2020-0 | ···················· | ···················· | ····················· | ···················· | ···················· | ···················· |
| W | 5124 hCoV-19/USA/AZ-TG322725/2020\|EPI_ISL_694336\|2020-0 | ···················· | ···················· | ····················· | ···················· | ···················· | ···················· |
| W | 5125 hCoV-19/USA/AZ-TG324558/2020\|EPI_ISL_694340\|2020-0 | ···················· | ···················· | ····················· | ···················· | ···················· | ···················· |
| W | 5126 hCoV-19/USA/AZ-TG324812/2020\|EPI_ISL_694341\|2020-0 | ···················· | ···················· | ····················· | ···················· | ···················· | ···················· |
| W | 5127 hCoV-19/USA/AZ-TG328534/2020\|EPI_ISL_694344\|2020-0 | ···················· | ·A·················· | ····················· | ···················· | ···················· | ···················· |
| W | 5128 hCoV-19/USA/AZ-TG329050/2020\|EPI_ISL_694347\|2020-0 | ···················· | ···················· | ····················· | ···················· | ···················· | ···················· |
| W | 5129 hCoV-19/USA/AZ-TG329100/2020\|EPI_ISL_694348\|2020-0 | ···················· | ···················· | ····················· | ···················· | ···················· | ···················· |
| W | 5130 hCoV-19/USA/AZ-TG329102/2020\|EPI_ISL_694349\|2020-0 | ···················· | ···················· | ····················· | ···················· | ···················· | ···················· |
| W | 5131 hCoV-19/USA/AZ-TG331333/2020\|EPI_ISL_694350\|2020-0 | ···················· | ···················· | ····················· | ···················· | ···················· | ···················· |
| W | 5132 hCoV-19/USA/AZ-TG331398/2020\|EPI_ISL_694351\|2020-0 | ···················· | ···················· | ····················· | ···················· | ···················· | ···················· |
| W | 5133 hCoV-19/USA/AZ-TG332555/2020\|EPI_ISL_694354\|2020-0 | ···················· | ···················· | ····················· | ···················· | ···················· | ···················· |
| W | 5134 hCoV-19/USA/AZ-TG603452/2020\|EPI_ISL_695656\|2020-0 | ···················· | ···················· | ····················· | ···················· | ···················· | ···················· |
| W | 5135 hCoV-19/USA/AZ-TG614806/2020\|EPI_ISL_695663\|2020-0 | ···················· | ·A·················· | ····················· | ···················· | ···················· | ···················· |
| W | 5136 hCoV-19/USA/AZ-TG616593/2020\|EPI_ISL_695664\|2020-0 | ···················· | ···················· | ····················· | ···················· | ···················· | ···················· |
| W | 5137 hCoV-19/USA/AZ-TG618007/2020\|EPI_ISL_695665\|2020-0 | ···················· | ···················· | ····················· | ···················· | ···················· | ···················· |
| W | 5138 hCoV-19/USA/AZ-TG618012/2020\|EPI_ISL_695667\|2020-0 | ···················· | ···················· | ····················· | ···················· | ···················· | ···················· |
| W | 5139 hCoV-19/USA/AZ-TG618024/2020\|EPI_ISL_695672\|2020-0 | ···················· | ···················· | ····················· | ···················· | ···················· | ···················· |
| W | 5140 hCoV-19/USA/AZ-TG618043/2020\|EPI_ISL_695685\|2020-0 | ···················· | ···················· | ····················· | ···················· | ···················· | ···················· |
| W | 5141 hCoV-19/United Arab Emirates/0426/2020\|EPI_ISL_698 | ···················· | ···················· | ····················· | ···················· | ···················· | ···················· |
| W | 5142 hCoV-19/United Arab Emirates/0964/2020\|EPI_ISL_699 | ···················· | ···················· | ····················· | ···················· | ···················· | ···················· |
| W | 5143 hCoV-19/United Arab Emirates/0965/2020\|EPI_ISL_699 | ···················· | ···················· | ····················· | ···················· | ···················· | ···················· |
| W | 5144 hCoV-19/United Arab Emirates/0966/2020\|EPI_ISL_699 | ···················· | ···················· | ····················· | ···················· | ···················· | ···················· |
| W | 5145 hCoV-19/United Arab Emirates/0967/2020\|EPI_ISL_699 | ···················· | ···················· | ····················· | ···················· | ···················· | ···················· |
| W | 5146 hCoV-19/United Arab Emirates/0968/2020\|EPI_ISL_699 | ···················· | ···················· | ····················· | ···················· | ···················· | ···················· |
| W | 5147 hCoV-19/United Arab Emirates/0969/2020\|EPI_ISL_699 | ···················· | ···················· | ····················· | ···················· | ···················· | ···················· |
| W | 5148 hCoV-19/United Arab Emirates/0970/2020\|EPI_ISL_699 | ···················· | ···················· | ····················· | ···················· | ···················· | ···················· |
| W | 5149 hCoV-19/United Arab Emirates/0972/2020\|EPI_ISL_699 | ···················· | ···················· | ····················· | ···················· | ···················· | ···················· |
| W | 5150 hCoV-19/United Arab Emirates/0973/2020\|EPI_ISL_699 | ···················· | ···················· | ····················· | ···················· | ···················· | ···················· |
| W | 5151 hCoV-19/United Arab Emirates/0974/2020\|EPI_ISL_699 | ···················· | ···················· | ····················· | ···················· | ···················· | ···················· |
| W | 5152 hCoV-19/United Arab Emirates/0975/2020\|EPI_ISL_699 | ···················· | ···················· | ····················· | ···················· | ···················· | ···················· |
| W | 5153 hCoV-19/United Arab Emirates/0976/2020\|EPI_ISL_699 | ···················· | ·A·················· | ····················· | ···················· | ···················· | ···················· |
| W | 5154 hCoV-19/United Arab Emirates/0977/2020\|EPI_ISL_699 | ···················· | ···················· | ····················· | ···················· | ···················· | ···················· |
| W | 5155 hCoV-19/United Arab Emirates/0978/2020\|EPI_ISL_699 | ···················· | ···················· | ····················· | ···················· | ···················· | ···················· |
| W | 5156 hCoV-19/United Arab Emirates/0980/2020\|EPI_ISL_699 | ···················· | ···················· | ····················· | ···················· | ···················· | ···················· |
| W | 5157 hCoV-19/United Arab Emirates/0981/2020\|EPI_ISL_699 | ···················· | ···················· | ····················· | ···················· | ···················· | ···················· |
| W | 5158 hCoV-19/United Arab Emirates/0982/2020\|EPI_ISL_699 | ···················· | ···················· | ····················· | ···················· | ···················· | ···················· |
| W | 5159 hCoV-19/United Arab Emirates/0983/2020\|EPI_ISL_699 | ···················· | ···················· | ····················· | ···················· | ···················· | ···················· |
| W | 5160 hCoV-19/United Arab Emirates/0984/2020\|EPI_ISL_699 | ···················· | ···················· | ····················· | ···················· | ···················· | ···················· |
| W | 5161 hCoV-19/United Arab Emirates/0986/2020\|EPI_ISL_699 | ···················· | ···················· | ····················· | ···················· | ···················· | ···················· |
| W | 5162 hCoV-19/United Arab Emirates/0987/2020\|EPI_ISL_699 | ···················· | ···················· | ····················· | ···················· | ···················· | ···················· |
| W | 5163 hCoV-19/United Arab Emirates/0988/2020\|EPI_ISL_699 | ···················· | ···················· | ····················· | ···················· | ···················· | ···················· |
| W | 5164 hCoV-19/United Arab Emirates/0989/2020\|EPI_ISL_699 | ···················· | ···················· | ····················· | ···················· | ···················· | ···················· |
| W | 5165 hCoV-19/United Arab Emirates/0990/2020\|EPI_ISL_699 | ···················· | ···················· | ····················· | ···················· | ···················· | ···················· |
| W | 5166 hCoV-19/United Arab Emirates/0991/2020\|EPI_ISL_699 | ···················· | ···················· | ····················· | ···················· | ···················· | ···················· |
| W | 5167 hCoV-19/United Arab Emirates/0992/2020\|EPI_ISL_699 | ···················· | ···················· | ····················· | ···················· | ···················· | ···················· |
| W | 5168 hCoV-19/United Arab Emirates/0993/2020\|EPI_ISL_699 | ···················· | ···················· | ····················· | ···················· | ···················· | ···················· |

| | | | | | | | | |
|---|---|---|---|---|---|---|---|---|
| W | 5169 hCoV-19/United Arab Emirates/0994/2020\|EPI_ISL_699 | .................... | .................... | .................... | .................... | .................... | .................... | |
| W | 5170 hCoV-19/United Arab Emirates/0996/2020\|EPI_ISL_699 | .................... | .................... | .................... | .................... | .................... | .................... | |
| W | 5171 hCoV-19/United Arab Emirates/0997/2020\|EPI_ISL_699 | .................... | .A.................. | .................... | .................... | .................... | .................... | |
| W | 5172 hCoV-19/United Arab Emirates/0998/2020\|EPI_ISL_699 | .................... | .................... | .................... | .................... | .................... | .................... | |
| W | 5173 hCoV-19/United Arab Emirates/0999/2020\|EPI_ISL_699 | .................... | .................... | .................... | .................... | .................... | .................... | |
| W | 5174 hCoV-19/United Arab Emirates/1000/2020\|EPI_ISL_699 | .................... | .................... | .................... | .................... | .................... | .................... | |
| W | 5175 hCoV-19/United Arab Emirates/1001/2020\|EPI_ISL_699 | .................... | .................... | .................... | .................... | .................... | .................... | |
| W | 5176 hCoV-19/United Arab Emirates/1002/2020\|EPI_ISL_699 | .................... | .................... | .................... | .................... | .................... | .................... | |
| W | 5177 hCoV-19/United Arab Emirates/1003/2020\|EPI_ISL_699 | .................... | .................... | .................... | .................... | .................... | .................... | |
| W | 5178 hCoV-19/United Arab Emirates/1004/2020\|EPI_ISL_699 | .................... | .................... | .................... | .................... | .................... | .................... | |
| W | 5179 hCoV-19/United Arab Emirates/1005/2020\|EPI_ISL_699 | .................... | .................... | .................... | .................... | .................... | .................... | |
| W | 5180 hCoV-19/United Arab Emirates/1006/2020\|EPI_ISL_699 | .................... | .................... | .................... | .................... | .................... | .................... | |
| W | 5181 hCoV-19/United Arab Emirates/1007/2020\|EPI_ISL_699 | .................... | .................... | .................... | .................... | .................... | .................... | |
| W | 5182 hCoV-19/United Arab Emirates/1008/2020\|EPI_ISL_699 | .................... | .................... | .................... | .................... | .................... | .................... | |
| W | 5183 hCoV-19/United Arab Emirates/1009/2020\|EPI_ISL_699 | .................... | .................... | .................... | .................... | .................... | .................... | |
| W | 5184 hCoV-19/United Arab Emirates/1010/2020\|EPI_ISL_699 | .................... | .................... | .................... | .................... | .................... | .................... | |
| W | 5185 hCoV-19/United Arab Emirates/1011/2020\|EPI_ISL_699 | .................... | .................... | .................... | .................... | .................... | .................... | |
| W | 5186 hCoV-19/United Arab Emirates/1012/2020\|EPI_ISL_699 | .................... | .................... | .................... | .................... | .................... | .................... | |
| W | 5187 hCoV-19/United Arab Emirates/1013/2020\|EPI_ISL_699 | .................... | .................... | .................... | .................... | .................... | .................... | |
| W | 5188 hCoV-19/United Arab Emirates/1014/2020\|EPI_ISL_699 | .................... | .................... | .................... | .................... | .................... | .................... | |
| W | 5189 hCoV-19/United Arab Emirates/1015/2020\|EPI_ISL_699 | .................... | .................... | .................... | .................... | .................... | .................... | |
| W | 5190 hCoV-19/United Arab Emirates/1016/2020\|EPI_ISL_699 | .................... | .................... | .................... | .................... | .................... | .................... | |
| W | 5191 hCoV-19/United Arab Emirates/1017/2020\|EPI_ISL_699 | .................... | .................... | .................... | .................... | .................... | .................... | |
| W | 5192 hCoV-19/United Arab Emirates/1018/2020\|EPI_ISL_699 | .................... | .................... | .................... | .................... | .................... | .................... | |
| W | 5193 hCoV-19/United Arab Emirates/1019/2020\|EPI_ISL_699 | .................... | .................... | .................... | .................... | .................... | .................... | |
| W | 5194 hCoV-19/United Arab Emirates/1020/2020\|EPI_ISL_699 | .................... | .................... | .................... | .................... | .................... | .................... | |
| W | 5195 hCoV-19/United Arab Emirates/1021/2020\|EPI_ISL_699 | .................... | .................... | .................... | .................... | .................... | .................... | |
| W | 5196 hCoV-19/United Arab Emirates/1022/2020\|EPI_ISL_699 | .................... | .................... | .................... | .................... | .................... | .................... | |
| W | 5197 hCoV-19/United Arab Emirates/1023/2020\|EPI_ISL_699 | .................... | .................... | .................... | .................... | .................... | .................... | |
| W | 5198 hCoV-19/United Arab Emirates/1024/2020\|EPI_ISL_699 | .................... | .................... | .................... | .................... | .................... | .................... | |
| W | 5199 hCoV-19/United Arab Emirates/1025/2020\|EPI_ISL_699 | .................... | .................... | .................... | .................... | .................... | .................... | |
| W | 5200 hCoV-19/United Arab Emirates/1026/2020\|EPI_ISL_699 | .................... | .................... | .................... | .................... | .................... | .................... | |
| W | 5201 hCoV-19/United Arab Emirates/1027/2020\|EPI_ISL_699 | .................... | .................... | .................... | .................... | .................... | .................... | |
| W | 5202 hCoV-19/United Arab Emirates/1028/2020\|EPI_ISL_699 | .................... | .................... | .................... | .................... | .................... | .................... | |
| W | 5203 hCoV-19/United Arab Emirates/1029/2020\|EPI_ISL_699 | .................... | .................... | .................... | .................... | .................... | .................... | |
| W | 5204 hCoV-19/India/MH-ACTREC-269/2020\|EPI_ISL_699921\|20 | .................... | .................... | .................... | .................... | .................... | .................... | |
| W | 5205 hCoV-19/India/MH-ACTREC-270/2020\|EPI_ISL_699922\|20 | .................... | .................... | .................... | .................... | .................... | .................... | |
| W | 5207 hCoV-19/India/MH-ACTREC-272/2020\|EPI_ISL_699924\|20 | .................... | .................... | .................... | .................... | .................... | .................... | |
| W | 5208 hCoV-19/India/MH-ACTREC-274/2020\|EPI_ISL_699926\|20 | .................... | .................... | .................... | .................... | .................... | .................... | |
| W | 5209 hCoV-19/India/MH-ACTREC-275/2020\|EPI_ISL_699927\|20 | .................... | .................... | .................... | .................... | .................... | .................... | |
| W | 5210 hCoV-19/India/MH-ACTREC-276/2020\|EPI_ISL_699928\|20 | .................... | .................... | .................... | .................... | .................... | .................... | |
| W | 5211 hCoV-19/India/MH-ACTREC-277/2020\|EPI_ISL_699929\|20 | .................... | .................... | .................... | .................... | .................... | .................... | |
| W | 5212 hCoV-19/India/MH-ACTREC-278/2020\|EPI_ISL_699930\|20 | .................... | .................... | .................... | .................... | .................... | .................... | |
| W | 5213 hCoV-19/India/MH-ACTREC-279/2020\|EPI_ISL_699931\|20 | .................... | .................... | .................... | .................... | .................... | .................... | |
| W | 5214 hCoV-19/England/CAMC-BD1331/2020\|EPI_ISL_702305\|20 | .................... | .................... | .................... | .................... | .................... | .................... | |
| W | 5215 hCoV-19/England/CAMC-BD1805/2020\|EPI_ISL_702306\|20 | .................... | .................... | .................... | .................... | .................... | .................... | |
| W | 5216 hCoV-19/England/CAMC-BD1EE5/2020\|EPI_ISL_702311\|20 | .................... | .A.................. | .................... | .................... | .................... | ..A................. | |
| W | 5217 hCoV-19/England/CAMC-BD1683/2020\|EPI_ISL_702320\|20 | .................... | .................... | .................... | .................... | .................... | .................... | |
| W | 5218 hCoV-19/England/CAMC-BD18D8/2020\|EPI_ISL_702322\|20 | .................... | .................... | .................... | .................... | .................... | .................... | |
| W | 5219 hCoV-19/England/CAMC-BD1A78/2020\|EPI_ISL_702324\|20 | .................... | .................... | .................... | .................... | .................... | .................... | |
| W | 5220 hCoV-19/England/CAMC-BD23B8/2020\|EPI_ISL_702325\|20 | .................... | .................... | .................... | .................... | .................... | .................... | |
| W | 5221 hCoV-19/England/CAMC-BD2154/2020\|EPI_ISL_702332\|20 | .................... | .................... | .................... | .................... | .....T.............. | .................... | |
| W | 5222 hCoV-19/England/CAMC-BD1B2A/2020\|EPI_ISL_702334\|20 | .................... | .................... | .................... | .................... | .................... | .................... | |
| W | 5223 hCoV-19/England/CAMC-BD147A/2020\|EPI_ISL_702336\|20 | .................... | .................... | .................... | .................... | .....T.............. | .................... | |
| W | 5224 hCoV-19/England/CAMC-BD1ED6/2020\|EPI_ISL_702341\|20 | .................... | .................... | .................... | .................... | .................... | .................... | |
| W | 5225 hCoV-19/England/CAMC-BD235E/2020\|EPI_ISL_702346\|20 | .................... | .................... | .................... | .................... | .................... | .................... | |
| W | 5226 hCoV-19/England/CAMC-BD1A69/2020\|EPI_ISL_702347\|20 | .................... | .................... | .................... | .................... | .................... | .................... | |
| W | 5227 hCoV-19/England/CAMC-BD1780/2020\|EPI_ISL_702348\|20 | .................... | .................... | .................... | .................... | .................... | .................... | |
| W | 5228 hCoV-19/England/CAMC-BD4459/2020\|EPI_ISL_702354\|20 | .................... | .................... | .................... | .................... | .................... | .................... | |
| W | 5229 hCoV-19/England/CAMC-BD2BED/2020\|EPI_ISL_702355\|20 | .................... | .................... | .................... | .................... | .................... | .................... | |
| W | 5230 hCoV-19/England/CAMC-BD435C/2020\|EPI_ISL_702356\|20 | .................... | .................... | .................... | .................... | .................... | .................... | |
| W | 5231 hCoV-19/England/CAMC-BD1B39/2020\|EPI_ISL_702357\|20 | .................... | .................... | .................... | .................... | .................... | .................... | |
| W | 5232 hCoV-19/England/CAMC-BD1F6A/2020\|EPI_ISL_702358\|20 | .................... | .................... | .................... | .................... | .....T.............. | .................... | |
| W | 5233 hCoV-19/England/CAMC-BD1F3D/2020\|EPI_ISL_702360\|20 | .................... | .................... | .................... | .................... | .................... | .................... | |
| W | 5234 hCoV-19/England/CAMC-BD15E0/2020\|EPI_ISL_702363\|20 | .................... | .................... | ...A................ | .................... | .................... | .................... | |
| W | 5235 hCoV-19/England/CAMC-BD1850/2020\|EPI_ISL_702364\|20 | .................... | .................... | .................... | .................... | .................... | .................... | |
| W | 5236 hCoV-19/England/CAMC-BD43B6/2020\|EPI_ISL_702367\|20 | .................... | .................... | .................... | .................... | .................... | .................... | |
| W | 5237 hCoV-19/England/CAMC-BD2303/2020\|EPI_ISL_702368\|20 | .................... | .................... | .................... | .................... | .................... | .................... | |
| W | 5238 hCoV-19/England/ALDP-BBAD7C/2020\|EPI_ISL_702370\|20 | .................... | .................... | .................... | .................... | .................... | .................... | |
| W | 5239 hCoV-19/Wales/ALDP-BBA468/2020\|EPI_ISL_702371\|2020 | ..................T. | .................... | .................... | .................... | .................... | .................... | 1 |
| W | 5240 hCoV-19/England/ALDP-BBA9F0/2020\|EPI_ISL_702372\|20 | .................... | .................... | .................... | .................... | .................... | .................... | |
| W | 5241 hCoV-19/England/ALDP-BB4E24/2020\|EPI_ISL_702373\|20 | .................... | .................... | .................... | .................... | .................... | .................... | |
| W | 5242 hCoV-19/England/ALDP-BB4923/2020\|EPI_ISL_702374\|20 | .................... | .................... | .................... | .................... | .....T.............. | .................... | |
| W | 5243 hCoV-19/England/ALDP-BB9BCE/2020\|EPI_ISL_702375\|20 | .................... | .................... | .................... | .................... | .................... | .................... | |
| W | 5244 hCoV-19/England/ALDP-BBA90F/2020\|EPI_ISL_702377\|20 | .................... | .................... | .................... | .................... | .................... | .................... | |
| W | 5245 hCoV-19/England/ALDP-BB96AF/2020\|EPI_ISL_702378\|20 | .................... | .................... | .................... | .................... | .................... | .................... | |
| W | 5246 hCoV-19/England/ALDP-BBA9B4/2020\|EPI_ISL_702379\|20 | .................... | .................... | .................... | .................... | .................... | .................... | |
| W | 5247 hCoV-19/England/ALDP-BB92C9/2020\|EPI_ISL_702382\|20 | .................... | .................... | .................... | .................... | .................... | .................... | |
| W | 5248 hCoV-19/England/ALDP-BB4A4E/2020\|EPI_ISL_702383\|20 | .................... | .................... | .................... | .................... | .................... | .................... | |
| W | 5249 hCoV-19/England/ALDP-BBAD5E/2020\|EPI_ISL_702387\|20 | .................... | .................... | .................... | .................... | .................... | .................... | |

| | | | | | | | | |
|---|---|---|---|---|---|---|---|---|
| W | 5250 | hCoV-19/England/ALDP-BB4F21/2020\|EPI_ISL_702388\|20 | ···················· | ···················· | ···················· | ···················· | ···················· | ···················· |
| W | 5251 | hCoV-19/England/ALDP-BB4E9D/2020\|EPI_ISL_702389\|20 | ···················· | ···················· | ···················· | ···················· | ···················· | ···················· |
| W | 5252 | hCoV-19/England/ALDP-BB9BDD/2020\|EPI_ISL_702390\|20 | ···················· | ···················· | ···················· | ···················· | ···················· | ···················· |
| W | 5253 | hCoV-19/England/ALDP-BBA459/2020\|EPI_ISL_702393\|20 | ···················· | ···················· | ···················· | ···················· | ···················· | ···················· |
| W | 5254 | hCoV-19/England/ALDP-BBA653/2020\|EPI_ISL_702394\|20 | ···················· | ·A·················· | ···················· | ···················· | ···················· | ···A················ |
| W | 5255 | hCoV-19/England/ALDP-BBA583/2020\|EPI_ISL_702395\|20 | ···················· | ···················· | ···················· | ···················· | ···················· | ···················· |
| W | 5256 | hCoV-19/England/ALDP-BB933F/2020\|EPI_ISL_702396\|20 | ···················· | ···················· | ···················· | ···················· | ···················· | ···················· |
| W | 5257 | hCoV-19/England/ALDP-BB92D8/2020\|EPI_ISL_702397\|20 | ···················· | ···················· | ···················· | ···················· | ···················· | ···················· |
| W | 5258 | hCoV-19/England/ALDP-BB4E8E/2020\|EPI_ISL_702398\|20 | ···················· | ···················· | ···················· | ···················· | ···················· | ···················· |
| W | 5259 | hCoV-19/England/ALDP-BBB555/2020\|EPI_ISL_702399\|20 | ···················· | ···················· | ···················· | ···················· | ···················· | ···················· |
| W | 5260 | hCoV-19/England/QEUH-BC3F2F/2020\|EPI_ISL_703600\|20 | ···················· | ···················· | ···················· | ···················· | ···················· | ···················· |
| W | 5261 | hCoV-19/England/ALDP-BCA562/2020\|EPI_ISL_703602\|20 | ···················· | ···················· | ···················· | ···················· | ···················· | ···················· |
| W | 5262 | hCoV-19/England/ALDP-BCA83C/2020\|EPI_ISL_703605\|20 | ···················· | ···················· | ···················· | ···················· | ···················· | ···················· |
| W | 5263 | hCoV-19/England/QEUH-BC47F9/2020\|EPI_ISL_703608\|20 | ···················· | ···················· | ···················· | ···················· | ···················· | ···················· |
| W | 5264 | hCoV-19/England/QEUH-BC4225/2020\|EPI_ISL_703612\|20 | ···················· | ···················· | ···················· | ···················· | ······T············· | ···················· |
| W | 5265 | hCoV-19/England/ALDP-BCA2B6/2020\|EPI_ISL_703613\|20 | ···················· | ···················· | ···················· | ···················· | ···················· | ···················· |
| W | 5266 | hCoV-19/England/ALDP-BCA7B7/2020\|EPI_ISL_703615\|20 | ···················· | ···················· | ···················· | ···················· | ···················· | ···················· |
| W | 5267 | hCoV-19/England/QEUH-BC4568/2020\|EPI_ISL_703617\|20 | ···················· | ···················· | ···················· | ···················· | ···················· | ···················· |
| W | 5268 | hCoV-19/England/ALDP-BCAA27/2020\|EPI_ISL_703618\|20 | ···················· | ···················· | ···················· | ···················· | ···················· | ···················· |
| W | 5269 | hCoV-19/England/ALDP-BCA92A/2020\|EPI_ISL_703620\|20 | ···················· | ···················· | ···················· | ···················· | ···················· | ···A················ |
| W | 5270 | hCoV-19/England/ALDP-BCA27A/2020\|EPI_ISL_703623\|20 | ···················· | ···················· | ···················· | ···················· | ···················· | ···················· |
| W | 5271 | hCoV-19/England/QEUH-BC486F/2020\|EPI_ISL_703626\|20 | ···················· | ···················· | ···················· | ···················· | ···················· | ···················· |
| W | 5272 | hCoV-19/England/ALDP-BCA289/2020\|EPI_ISL_703628\|20 | ···················· | ···················· | ···················· | ···················· | ···················· | ···················· |
| W | 5273 | hCoV-19/England/ALDP-BCA68D/2020\|EPI_ISL_703630\|20 | ···················· | ···················· | ···················· | ···················· | ···················· | ···················· |
| W | 5274 | hCoV-19/England/QEUH-BC4586/2020\|EPI_ISL_703631\|20 | ···················· | ···················· | ···················· | ···················· | ···················· | ···················· |
| W | 5275 | hCoV-19/England/QEUH-BC46CF/2020\|EPI_ISL_703633\|20 | ···················· | ···················· | ···················· | ···················· | ···················· | ···················· |
| W | 5276 | hCoV-19/England/QEUH-BC4674/2020\|EPI_ISL_703634\|20 | ···················· | ···················· | ···················· | ···················· | ···················· | ···················· |
| W | 5277 | hCoV-19/England/QEUH-BC4322/2020\|EPI_ISL_703640\|20 | ···················· | ···················· | ···················· | ···················· | ···················· | ···················· |
| W | 5278 | hCoV-19/England/QEUH-BC3FA7/2020\|EPI_ISL_703641\|20 | ···················· | ···················· | ···················· | ···················· | ···················· | ···················· |
| W | 5279 | hCoV-19/England/QEUH-BC4058/2020\|EPI_ISL_703645\|20 | ···················· | ···················· | ···················· | ···················· | ···················· | ···················· |
| W | 5280 | hCoV-19/England/ALDP-BCA131/2020\|EPI_ISL_703646\|20 | ···················· | ···················· | ···················· | ···················· | ···················· | ···················· |
| W | 5281 | hCoV-19/England/ALDP-BCA4DE/2020\|EPI_ISL_703648\|20 | ···················· | ···················· | ···················· | ···················· | ···················· | ···················· |
| W | 5282 | hCoV-19/England/QEUH-BC4067/2020\|EPI_ISL_703650\|20 | ···················· | ···················· | ···················· | ···················· | ···················· | ···················· |
| W | 5283 | hCoV-19/England/QEUH-BC46ED/2020\|EPI_ISL_703651\|20 | ···················· | ···················· | ···················· | ···················· | ···················· | ···················· |
| W | 5284 | hCoV-19/England/QEUH-BC461A/2020\|EPI_ISL_703657\|20 | ···················· | ···················· | ···················· | ···················· | ······T············· | ···················· |
| W | 5285 | hCoV-19/England/QEUH-BC4128/2020\|EPI_ISL_703659\|20 | ···················· | ···················· | ···················· | ···················· | ···················· | ···················· |
| W | 5286 | hCoV-19/England/QEUH-BC48AB/2020\|EPI_ISL_703661\|20 | ···················· | ···················· | ···················· | ···················· | ···················· | ···················· |
| W | 5287 | hCoV-19/England/ALDP-BCA31D/2020\|EPI_ISL_703662\|20 | ···················· | ···················· | ···················· | ···················· | ···················· | ···················· |
| W | 5288 | hCoV-19/England/ALDP-BCA465/2020\|EPI_ISL_703664\|20 | ···················· | ···················· | ···················· | ···················· | ···················· | ···················· |
| W | 5289 | hCoV-19/England/ALDP-BCA984/2020\|EPI_ISL_703665\|20 | ···················· | ·········T·········· | ···················· | ···················· | ···················· | ···················· |
| W | 5290 | hCoV-19/England/QEUH-BC44D4/2020\|EPI_ISL_703672\|20 | ···················· | ···················· | ···················· | ···················· | ···················· | ···················· |
| W | 5291 | hCoV-19/England/QEUH-BC45B3/2020\|EPI_ISL_703673\|20 | ···················· | ···················· | ···················· | ···················· | ···················· | ···················· |
| W | 5292 | hCoV-19/England/ALDP-BCA8F0/2020\|EPI_ISL_703675\|20 | ···················· | ···················· | ···················· | ···················· | ···················· | ···················· |
| W | 5293 | hCoV-19/England/QEUH-BC4489/2020\|EPI_ISL_703677\|20 | ···················· | ···················· | ···················· | ···················· | ···················· | ···················· |
| W | 5294 | hCoV-19/England/ALDP-BCA8C3/2020\|EPI_ISL_703678\|20 | ···················· | ···················· | ···················· | ···················· | ···················· | ···················· |
| W | 5295 | hCoV-19/England/ALDP-BCA9FD/2020\|EPI_ISL_703681\|20 | ···················· | ···················· | ···················· | ···················· | ···················· | ···················· |
| W | 5296 | hCoV-19/England/QEUH-BC447A/2020\|EPI_ISL_703685\|20 | ···················· | ···················· | ···················· | ···················· | ···················· | ···················· |
| W | 5297 | hCoV-19/England/ALDP-BCA2C5/2020\|EPI_ISL_703686\|20 | ···················· | ···················· | ···················· | ···················· | ···················· | ···················· |
| W | 5298 | hCoV-19/England/ALDP-BCA6AB/2020\|EPI_ISL_703688\|20 | ···················· | ···················· | ···················· | ···················· | ···················· | ···················· |
| W | 5299 | hCoV-19/England/ALDP-BCA5F9/2020\|EPI_ISL_703691\|20 | ···················· | ···················· | ···················· | ···················· | ···················· | ···················· |
| W | 5300 | hCoV-19/England/ALDP-BCA492/2020\|EPI_ISL_703692\|20 | ···················· | ···················· | ···················· | ···················· | ···················· | ···················· |
| W | 5301 | hCoV-19/England/QEUH-BC48F6/2020\|EPI_ISL_703694\|20 | ···················· | ···················· | ···················· | ···················· | ···················· | ···················· |
| W | 5302 | hCoV-19/England/ALDP-BCA8D2/2020\|EPI_ISL_703696\|20 | ···················· | ···················· | ···················· | ···················· | ···················· | ···················· |
| W | 5303 | hCoV-19/England/ALDP-BCA948/2020\|EPI_ISL_703699\|20 | ···················· | ···················· | ···················· | ···················· | ···················· | ···················· |
| W | 5304 | hCoV-19/England/QEUH-BC4A2D/2020\|EPI_ISL_704130\|20 | ···················· | ···················· | ···················· | ···················· | ···················· | ···················· |
| W | 5305 | hCoV-19/England/QEUH-BC39D6/2020\|EPI_ISL_704133\|20 | ···················· | ···················· | ···················· | ···················· | ···················· | ···················· |
| W | 5306 | hCoV-19/England/QEUH-BC4AA5/2020\|EPI_ISL_704135\|20 | ···················· | ···················· | ···················· | ···················· | ···················· | ···················· |
| W | 5307 | hCoV-19/England/QEUH-BCB9FC/2020\|EPI_ISL_704136\|20 | ···················· | ···················· | ···················· | ···················· | ···················· | ···················· |
| W | 5308 | hCoV-19/England/CAMC-BC01B3/2020\|EPI_ISL_704902\|20 | ···················· | ·A·················· | ···················· | ···················· | ···················· | ···A················ |
| W | 5309 | hCoV-19/England/LIVE-DC19B8/2020\|EPI_ISL_704903\|20 | ···················· | ···················· | ···················· | ···················· | ···················· | ···················· |
| W | 5310 | hCoV-19/England/CAMC-BC0933/2020\|EPI_ISL_704904\|20 | ···················· | ···················· | ···················· | ···················· | ···················· | ···················· |
| W | 5311 | hCoV-19/England/LIVE-DC19C7/2020\|EPI_ISL_704905\|20 | ···················· | ···················· | ···················· | ···················· | ···················· | ···················· |
| W | 5312 | hCoV-19/England/CAMC-BC0F7D/2020\|EPI_ISL_704906\|20 | ···················· | ···················· | ···················· | ···················· | ···················· | ···················· |
| W | 5313 | hCoV-19/England/CAMC-BC002F/2020\|EPI_ISL_704907\|20 | ···················· | ···················· | ···················· | ···················· | ···················· | ···················· |
| W | 5314 | hCoV-19/England/NORW-EED89/2020\|EPI_ISL_704908\|202 | ···················· | ·A·················· | ···················· | ···················· | ···················· | ···A················ |
| W | 5315 | hCoV-19/England/NORW-F778F/2020\|EPI_ISL_704911\|202 | ···················· | ·A·················· | ···················· | ···················· | ···················· | ···A················ |
| W | 5316 | hCoV-19/England/CAMC-BC0B6A/2020\|EPI_ISL_704912\|20 | ···················· | ···················· | ···················· | ···················· | ···················· | ···················· |
| W | 5317 | hCoV-19/England/NORW-F8B37/2020\|EPI_ISL_704914\|202 | ···················· | ·A·················· | ···················· | ···················· | ···················· | ···A················ |
| W | 5318 | hCoV-19/England/NORW-F918F/2020\|EPI_ISL_704916\|202 | ···················· | ·A·················· | ···················· | ···················· | ···················· | ···A················ |
| W | 5319 | hCoV-19/England/CAMC-BC07EE/2020\|EPI_ISL_704917\|20 | ···················· | ···················· | ···················· | ···················· | ···················· | ···················· |
| W | 5321 | hCoV-19/England/NORW-F6586/2020\|EPI_ISL_704921\|202 | ···················· | ···················· | ···················· | ···················· | ···················· | ···················· |
| W | 5322 | hCoV-19/England/CAMC-BC03EA/2020\|EPI_ISL_704922\|20 | ···················· | ·A·················· | ···················· | ···················· | ···················· | ···A················ |
| W | 5323 | hCoV-19/England/NORW-F97F6/2020\|EPI_ISL_704923\|202 | ···················· | ···················· | ···················· | ···················· | ···················· | ···················· |
| W | 5324 | hCoV-19/England/NORW-F9820/2020\|EPI_ISL_704926\|202 | ···················· | ···················· | ···················· | ···················· | ···················· | ···················· |
| W | 5325 | hCoV-19/England/CAMC-BBFF35/2020\|EPI_ISL_704927\|20 | ···················· | ···················· | ···················· | ···················· | ···················· | ···················· |
| W | 5326 | hCoV-19/England/CAMC-BC08FA/2020\|EPI_ISL_704928\|20 | ···················· | ···················· | ···················· | ···················· | ···················· | ···················· |
| W | 5327 | hCoV-19/England/NORW-F984E/2020\|EPI_ISL_704929\|202 | ···················· | ···················· | ···················· | ···················· | ···················· | ···················· |
| W | 5328 | hCoV-19/England/CAMC-BC0317/2020\|EPI_ISL_704935\|20 | ···················· | ···················· | ···················· | ···················· | ···················· | ···················· |
| W | 5329 | hCoV-19/England/CAMC-BC0DDD/2020\|EPI_ISL_704938\|20 | ···················· | ···················· | ···················· | ···················· | ···················· | ···················· |
| W | 5330 | hCoV-19/England/CAMC-BC0E34/2020\|EPI_ISL_704943\|20 | ···················· | ···················· | ···················· | ···················· | ···················· | ···················· |

W 5331 hCoV-19/England/CAMC-BC0B2E/2020|EPI_ISL_704946|20 ···················· ···················· ···················· ···················· ····T··············· ····················
W 5332 hCoV-19/England/CAMC-BC1C48/2020|EPI_ISL_704949|20 ···················· ···················· ···················· ···················· ···················· ····················
W 5333 hCoV-19/England/CAMC-BC0292/2020|EPI_ISL_704951|20 ···················· ···················· ···················· ···················· ···················· ····················
W 5334 hCoV-19/England/CAMC-BC0B1F/2020|EPI_ISL_704953|20 ···················· ···················· ···················· ···················· ···················· ····················
W 5335 hCoV-19/England/CAMC-BC00E3/2020|EPI_ISL_704956|20 ···················· ···················· ···················· ···················· ······T············· ····················
W 5336 hCoV-19/England/CAMC-BC0AE5/2020|EPI_ISL_704958|20 ···················· ···················· ···················· ···················· ···················· ····················
W 5337 hCoV-19/England/CAMC-BC0168/2020|EPI_ISL_704961|20 ········T··········· ···················· ···················· ···················· ···················· ····················
W 5338 hCoV-19/England/CAMC-BC0511/2020|EPI_ISL_704962|20 ···················· ······A············· ···················· ···················· ···················· ····················
W 5339 hCoV-19/England/CAMC-BC0380/2020|EPI_ISL_704974|20 ···················· ···················· ···················· ···················· ······T············· ····················
W 5340 hCoV-19/England/CAMC-BC0414/2020|EPI_ISL_704975|20 ···················· ···················· ···················· ···················· ···················· ····················
W 5341 hCoV-19/England/CAMC-BC1EE8/2020|EPI_ISL_704979|20 ···················· ·A·················· ···················· ···················· ···················· ···A················
W 5342 hCoV-19/England/CAMC-BC0A4F/2020|EPI_ISL_704980|20 ···················· ···················· ···················· ···················· ···················· ····················
W 5343 hCoV-19/England/CAMC-BC07DF/2020|EPI_ISL_704984|20 ···················· ···················· ···················· ···················· ···················· ····················
W 5344 hCoV-19/England/CAMC-BBFEFC/2020|EPI_ISL_704987|20 ···················· ·A·················· ···················· ···················· ···················· ····················
W 5345 hCoV-19/England/CAMC-BC006B/2020|EPI_ISL_704988|20 ···················· ···················· ···················· ···················· ···················· ····················
W 5346 hCoV-19/England/CAMC-BC20D2/2020|EPI_ISL_704992|20 ···················· ···················· ···················· ···················· ···················· ····················
W 5347 hCoV-19/England/CAMC-BC1A20/2020|EPI_ISL_704993|20 ···················· ···················· ···················· ···················· ······T············· ····················
W 5348 hCoV-19/Wales/PHWC-491EAC/2020|EPI_ISL_706790|2020 ···················· ···················· ···················· ···················· ···················· ····················
W 5353 hCoV-19/Wales/PHWC-491EF7/2020|EPI_ISL_706795|2020 ···················· ···················· ···················· ···················· ···················· ····················
W 5356 hCoV-19/Wales/PHWC-491F5E/2020|EPI_ISL_706798|2020 ···················· ···················· ···················· ···················· ···················· ····················
W 5357 hCoV-19/USA/MN-MDH-2132/2020|EPI_ISL_708470|2020-1 ···················· ···················· ···················· ···················· ···················· ····················
W 5358 hCoV-19/USA/MN-MDH-2133/2020|EPI_ISL_708471|2020-1 ···················· ···················· ···················· ···················· ···················· ····················
W 5359 hCoV-19/USA/MN-MDH-2134/2020|EPI_ISL_708472|2020-1 ···················· ···················· ···················· ···················· ···················· ····················
W 5360 hCoV-19/USA/MN-MDH-2135/2020|EPI_ISL_708473|2020-1 ···················· ···················· ···················· ···················· ···················· ····················
W 5361 hCoV-19/USA/MN-MDH-2136/2020|EPI_ISL_708474|2020-1 ···················· ···················· ···················· ···················· ···················· ····················
W 5362 hCoV-19/USA/MN-MDH-2137/2020|EPI_ISL_708475|2020-1 ···················· ···················· ···················· ···················· ···················· ····················
W 5363 hCoV-19/USA/MN-MDH-2138/2020|EPI_ISL_708476|2020-1 ···················· ···················· ···················· ···················· ···················· ····················
W 5364 hCoV-19/USA/MN-MDH-2139/2020|EPI_ISL_708477|2020-1 ···················· ···················· ···················· ···················· ···················· ····················
W 5365 hCoV-19/USA/MN-MDH-2140/2020|EPI_ISL_708478|2020-1 ···················· ···················· ···················· ···················· ···················· ····················
W 5366 hCoV-19/USA/MN-MDH-2141/2020|EPI_ISL_708479|2020-1 ···················· ···················· ···················· ···················· ···················· ····················
W 5367 hCoV-19/USA/MN-MDH-2142/2020|EPI_ISL_708480|2020-1 ···················· ·A·················· ···················· ···················· ···················· ····················
W 5368 hCoV-19/USA/MN-MDH-2143/2020|EPI_ISL_708481|2020-1 ···················· ···················· ···················· ···················· ···················· ····················
W 5369 hCoV-19/USA/MN-MDH-2144/2020|EPI_ISL_708482|2020-1 ···················· ·A·················· ···················· ···················· ···················· ····················
W 5370 hCoV-19/USA/MN-MDH-2145/2020|EPI_ISL_708483|2020-1 ···················· ···················· ···················· ···················· ···················· ····················
W 5371 hCoV-19/USA/MN-MDH-2146/2020|EPI_ISL_708484|2020-1 ···················· ·A·················· ···················· ···················· ···················· ····················
W 5372 hCoV-19/USA/MN-MDH-2147/2020|EPI_ISL_708485|2020-1 ···················· ···················· ···················· ···················· ···················· ····················
W 5373 hCoV-19/USA/MN-MDH-2148/2020|EPI_ISL_708486|2020-1 ···················· ···················· ···················· ···················· ···················· ····················
W 5374 hCoV-19/USA/MN-MDH-2149/2020|EPI_ISL_708487|2020-1 ···················· ·A·················· ···················· ···················· ···················· ····················
W 5375 hCoV-19/USA/MN-MDH-2150/2020|EPI_ISL_708488|2020-1 ···················· ···················· ···················· ···················· ···················· ····················
W 5376 hCoV-19/USA/MN-MDH-2151/2020|EPI_ISL_708489|2020-1 ···················· ···················· ···················· ···················· ···················· ····················
W 5377 hCoV-19/USA/AZ-TG654363/2020|EPI_ISL_696000|2020-0 ···················· ···················· ···················· ···················· ···················· ····················
W 5378 hCoV-19/USA/AZ-TG654368/2020|EPI_ISL_696005|2020-0 ···················· ···················· ···················· ···················· ···················· ····················
W 5379 hCoV-19/USA/AZ-TG654371/2020|EPI_ISL_696008|2020-0 ···················· ···················· ···················· ···················· ···················· ····················
W 5380 hCoV-19/USA/AZ-TG654694/2020|EPI_ISL_696015|2020-0 ···················· ···················· ···················· ···················· ···················· ····················
W 5381 hCoV-19/United Arab Emirates/0427/2020|EPI_ISL_698 ···················· ···················· ···················· ···················· ···················· ····················
W 5382 hCoV-19/United Arab Emirates/0428/2020|EPI_ISL_698 ···················· ···················· ···················· ···················· ···················· ····················
W 5383 hCoV-19/United Arab Emirates/0429/2020|EPI_ISL_698 ···················· ···················· ···················· ···················· ···················· ····················
W 5384 hCoV-19/United Arab Emirates/0430/2020|EPI_ISL_698 ···················· ···················· ···················· ···················· ···················· ····················
W 5385 hCoV-19/United Arab Emirates/0431/2020|EPI_ISL_698 ···················· ···················· ···················· ···················· ···················· ····················
W 5386 hCoV-19/United Arab Emirates/0432/2020|EPI_ISL_698 ···················· ···················· ···················· ···················· ···················· ····················
W 5387 hCoV-19/United Arab Emirates/0433/2020|EPI_ISL_698 ···················· ···················· ···················· ···················· ···················· ····················
W 5388 hCoV-19/United Arab Emirates/0434/2020|EPI_ISL_698 ···················· ···················· ···················· ···················· ···················· ····················
W 5389 hCoV-19/United Arab Emirates/0435/2020|EPI_ISL_698 ···················· ···················· ···················· ···················· ···················· ····················
W 5390 hCoV-19/United Arab Emirates/0436/2020|EPI_ISL_698 ···················· ···················· ···················· ···················· ···················· ····················
W 5391 hCoV-19/United Arab Emirates/0437/2020|EPI_ISL_698 ···················· ···················· ···················· ···················· ···················· ····················
W 5392 hCoV-19/United Arab Emirates/0438/2020|EPI_ISL_698 ···················· ···················· ···················· ···················· ···················· ····················
W 5393 hCoV-19/United Arab Emirates/0439/2020|EPI_ISL_698 ···················· ···················· ···················· ···················· ···················· ····················
W 5394 hCoV-19/United Arab Emirates/0440/2020|EPI_ISL_698 ···················· ···················· ···················· ···················· ···················· ····················
W 5395 hCoV-19/United Arab Emirates/0441/2020|EPI_ISL_698 ···················· ···················· ···················· ···················· ···················· ····················
W 5396 hCoV-19/United Arab Emirates/0442/2020|EPI_ISL_698 ···················· ···················· ···················· ···················· ···················· ····················
W 5397 hCoV-19/United Arab Emirates/0443/2020|EPI_ISL_698 ···················· ···················· ···················· ···················· ···················· ····················
W 5398 hCoV-19/United Arab Emirates/0445/2020|EPI_ISL_698 ···················· ···················· ···················· ···················· ···················· ····················
W 5399 hCoV-19/United Arab Emirates/0447/2020|EPI_ISL_698 ···················· ···················· ···················· ···················· ···················· ····················
W 5400 hCoV-19/United Arab Emirates/0448/2020|EPI_ISL_698 ···················· ···················· ···················· ···················· ···················· ····················
W 5401 hCoV-19/United Arab Emirates/0449/2020|EPI_ISL_698 ···················· ···················· ···················· ···················· ···················· ····················
W 5402 hCoV-19/United Arab Emirates/0451/2020|EPI_ISL_698 ···················· ···················· ···················· ···················· ···················· ····················
W 5403 hCoV-19/United Arab Emirates/0452/2020|EPI_ISL_698 ···················· ···················· ···················· ···················· ···················· ····················
W 5404 hCoV-19/United Arab Emirates/0453/2020|EPI_ISL_698 ···················· ···················· ···················· ···················· ···················· ····················
W 5405 hCoV-19/United Arab Emirates/0454/2020|EPI_ISL_698 ···················· ···················· ···················· ···················· ···················· ····················
W 5406 hCoV-19/United Arab Emirates/0456/2020|EPI_ISL_698 ···················· ···················· ···················· ···················· ···················· ····················
W 5407 hCoV-19/United Arab Emirates/0457/2020|EPI_ISL_698 ···················· ···················· ···················· ···················· ···················· ····················
W 5408 hCoV-19/United Arab Emirates/0458/2020|EPI_ISL_698 ···················· ···················· ···················· ···················· ···················· ····················
W 5409 hCoV-19/United Arab Emirates/0459/2020|EPI_ISL_698 ···················· ···················· ···················· ···················· ···················· ····················
W 5410 hCoV-19/United Arab Emirates/0460/2020|EPI_ISL_698 ···················· ···················· ···················· ···················· ···················· ····················
W 5411 hCoV-19/United Arab Emirates/0461/2020|EPI_ISL_698 ···················· ···················· ···················· ···················· ···················· ···············A····
W 5412 hCoV-19/United Arab Emirates/0462/2020|EPI_ISL_698 ···················· ···················· ···················· ···················· ···················· ····················
W 5413 hCoV-19/United Arab Emirates/0463/2020|EPI_ISL_698 ···················· ···················· ···················· ···················· ···················· ····················
W 5414 hCoV-19/United Arab Emirates/0464/2020|EPI_ISL_698 ···················· ···················· ···················· ···················· ···················· ····················
W 5415 hCoV-19/United Arab Emirates/0465/2020|EPI_ISL_698 ···················· ···················· ···················· ···················· ···················· ····················
W 5416 hCoV-19/United Arab Emirates/0466/2020|EPI_ISL_698 ···················· ···················· ···················· ···················· ···················· ····················

| | | | | | | | | |
|---|---|---|---|---|---|---|---|---|
| W | 5417 | hCoV-19/United Arab Emirates/0467/2020\|EPI_ISL_698 | ···················· | ···················· | ···················· | ···················· | ···················· | ···················· |
| W | 5418 | hCoV-19/United Arab Emirates/0468/2020\|EPI_ISL_698 | ···················· | ···················· | ···················· | ···················· | ···················· | ···················· |
| W | 5419 | hCoV-19/United Arab Emirates/0470/2020\|EPI_ISL_698 | ···················· | ···················· | ···················· | ···················· | ···T················ | ···················· |
| W | 5420 | hCoV-19/United Arab Emirates/0471/2020\|EPI_ISL_698 | ···················· | ···················· | ···················· | ···················· | ···················· | ···················· |
| W | 5421 | hCoV-19/United Arab Emirates/0472/2020\|EPI_ISL_698 | ···················· | ···················· | ···················· | ···················· | ···················· | ···················· |
| W | 5422 | hCoV-19/United Arab Emirates/0473/2020\|EPI_ISL_698 | ···················· | ···················· | ···················· | ···················· | ···················· | ···················· |
| W | 5423 | hCoV-19/United Arab Emirates/0474/2020\|EPI_ISL_698 | ···················· | ···················· | ···················· | ···················· | ···················· | ···················· |
| W | 5424 | hCoV-19/United Arab Emirates/0475/2020\|EPI_ISL_698 | ···················· | ···················· | ···················· | ···················· | ···················· | ···················· |
| W | 5425 | hCoV-19/United Arab Emirates/0476/2020\|EPI_ISL_698 | ···················· | ···················· | ···················· | ···················· | ···················· | ···················· |
| W | 5426 | hCoV-19/United Arab Emirates/0477/2020\|EPI_ISL_698 | ···················· | ···················· | ···················· | ···················· | ···················· | ···················· |
| W | 5427 | hCoV-19/United Arab Emirates/0478/2020\|EPI_ISL_698 | ···················· | ···················· | ···················· | ···················· | ···················· | ···················· |
| W | 5428 | hCoV-19/United Arab Emirates/0479/2020\|EPI_ISL_698 | ···················· | ···················· | ···················· | ···················· | ···················· | ···················· |
| W | 5429 | hCoV-19/United Arab Emirates/0481/2020\|EPI_ISL_698 | ···················· | ···················· | ···················· | ···················· | ···················· | ···················· |
| W | 5430 | hCoV-19/United Arab Emirates/0482/2020\|EPI_ISL_698 | ···················· | ···················· | ···········T········ | ···················· | ···················· | ···················· |
| W | 5431 | hCoV-19/United Arab Emirates/0483/2020\|EPI_ISL_698 | ···················· | ···················· | ···················· | ···················· | ···················· | ···················· |
| W | 5432 | hCoV-19/United Arab Emirates/0484/2020\|EPI_ISL_698 | ···················· | ···················· | ···················· | ···················· | ···················· | ···················· |
| W | 5433 | hCoV-19/United Arab Emirates/0485/2020\|EPI_ISL_698 | ···················· | ···················· | ···················· | ···················· | ···················· | ···················· |
| W | 5434 | hCoV-19/United Arab Emirates/0486/2020\|EPI_ISL_698 | ···················· | ···················· | ···················· | ···················· | ···················· | ···················· |
| W | 5435 | hCoV-19/United Arab Emirates/0487/2020\|EPI_ISL_698 | ···················· | ···················· | ···················· | ···················· | ···················· | ···················· |
| W | 5436 | hCoV-19/United Arab Emirates/0488/2020\|EPI_ISL_698 | ···················· | ···················· | ···················· | ···················· | ···················· | ···················· |
| W | 5437 | hCoV-19/United Arab Emirates/0489/2020\|EPI_ISL_698 | ···················· | ···················· | ···················· | ···················· | ···················· | ···················· |
| W | 5438 | hCoV-19/United Arab Emirates/0490/2020\|EPI_ISL_698 | ···················· | ···················· | ···················· | ···················· | ···················· | ···················· |
| W | 5439 | hCoV-19/United Arab Emirates/0492/2020\|EPI_ISL_698 | ···················· | ···················· | ···················· | ···················· | ···················· | ···················· |
| W | 5440 | hCoV-19/United Arab Emirates/0493/2020\|EPI_ISL_698 | ···················· | ···················· | ···················· | ···················· | ···················· | ···················· |
| W | 5441 | hCoV-19/United Arab Emirates/0494/2020\|EPI_ISL_698 | ···················· | ···················· | ···················· | ···················· | ···················· | ···················· |
| W | 5442 | hCoV-19/United Arab Emirates/0495/2020\|EPI_ISL_698 | ···················· | ···················· | ···················· | ···················· | ···················· | ···················· |
| W | 5443 | hCoV-19/United Arab Emirates/0496/2020\|EPI_ISL_698 | ···················· | ·A·················· | ···················· | ···················· | ···················· | ···················· |
| W | 5444 | hCoV-19/United Arab Emirates/0497/2020\|EPI_ISL_698 | ···················· | ···················· | ···················· | ···················· | ···················· | ···················· |
| W | 5445 | hCoV-19/United Arab Emirates/0498/2020\|EPI_ISL_698 | ···················· | ···················· | ···················· | ···················· | ···················· | ···················· |
| W | 5446 | hCoV-19/United Arab Emirates/0499/2020\|EPI_ISL_698 | ···················· | ···················· | ···················· | ···················· | ···················· | ···················· |
| W | 5447 | hCoV-19/United Arab Emirates/0500/2020\|EPI_ISL_698 | ···················· | ···················· | ···················· | ···················· | ···················· | ···················· |
| W | 5448 | hCoV-19/United Arab Emirates/0501/2020\|EPI_ISL_698 | ···················· | ···················· | ···················· | ···················· | ···················· | ···················· |
| W | 5449 | hCoV-19/United Arab Emirates/0502/2020\|EPI_ISL_698 | ···················· | ···················· | ···················· | ···················· | ···················· | ···················· |
| W | 5450 | hCoV-19/United Arab Emirates/0503/2020\|EPI_ISL_698 | ···················· | ···················· | ···················· | ···················· | ···················· | ···················· |
| W | 5451 | hCoV-19/United Arab Emirates/0504/2020\|EPI_ISL_698 | ···················· | ···················· | ···················· | ···················· | ···················· | ···················· |
| W | 5452 | hCoV-19/United Arab Emirates/0505/2020\|EPI_ISL_698 | ···················· | ···················· | ···················· | ···················· | ···················· | ···················· |
| W | 5453 | hCoV-19/United Arab Emirates/0507/2020\|EPI_ISL_698 | ···················· | ···················· | ···················· | ···················· | ···················· | ···················· |
| W | 5454 | hCoV-19/United Arab Emirates/0508/2020\|EPI_ISL_698 | ···················· | ···················· | ···················· | ···················· | ···················· | ···················· |
| W | 5455 | hCoV-19/United Arab Emirates/0509/2020\|EPI_ISL_698 | ···················· | ···················· | ···················· | ···················· | ···················· | ···················· |
| W | 5456 | hCoV-19/United Arab Emirates/0510/2020\|EPI_ISL_698 | ···················· | ···················· | ···················· | ···················· | ···················· | ···················· |
| W | 5457 | hCoV-19/United Arab Emirates/0511/2020\|EPI_ISL_698 | ···················· | ···················· | ···················· | ···················· | ···················· | ···················· |
| W | 5459 | hCoV-19/India/MH-ACTREC-281/2020\|EPI_ISL_699933\|20 | ···················· | ··········A········· | ···················· | ···················· | ···················· | ···················· |
| W | 5460 | hCoV-19/India/MH-ACTREC-282/2020\|EPI_ISL_699934\|20 | ···················· | ···················· | ···················· | ···················· | ···················· | ···················· |
| W | 5461 | hCoV-19/India/MH-ACTREC-283/2020\|EPI_ISL_699935\|20 | ···················· | ···················· | ···················· | ···················· | ···················· | ···················· |
| W | 5462 | hCoV-19/India/MH-ACTREC-284/2020\|EPI_ISL_699936\|20 | ···················· | ···················· | ···················· | ···················· | ···················· | ···················· |
| W | 5463 | hCoV-19/India/MH-ACTREC-285/2020\|EPI_ISL_699937\|20 | ···················· | ···················· | ···················· | ···················· | ···················· | ···················· |
| W | 5464 | hCoV-19/India/MH-ACTREC-286/2020\|EPI_ISL_699938\|20 | ···················· | ···················· | ···················· | ···················· | ···················· | ···················· |
| W | 5465 | hCoV-19/India/MH-ACTREC-287/2020\|EPI_ISL_699939\|20 | ···················· | ···················· | ···················· | ···················· | ···················· | ···················· |
| W | 5466 | hCoV-19/India/MH-ACTREC-289/2020\|EPI_ISL_699941\|20 | ···················· | ···················· | ···················· | ···················· | ···················· | ···················· |
| W | 5467 | hCoV-19/India/MH-ACTREC-290/2020\|EPI_ISL_699942\|20 | ···················· | ···················· | ···················· | ···················· | ···················· | ···················· |
| W | 5468 | hCoV-19/India/MH-ACTREC-291/2020\|EPI_ISL_699943\|20 | ···················· | ···················· | ···A················ | ···················· | ···················· | ···················· |
| W | 5469 | hCoV-19/India/MH-ACTREC-292/2020\|EPI_ISL_699944\|20 | ···················· | ···················· | ···················· | ···················· | ···················· | ···················· |
| W | 5470 | hCoV-19/India/MH-ACTREC-293/2020\|EPI_ISL_699945\|20 | ···················· | ···················· | ···················· | ···················· | ···················· | ···················· |
| W | 5471 | hCoV-19/India/MH-ACTREC-294/2020\|EPI_ISL_699946\|20 | ···················· | ···················· | ···················· | ···················· | ···················· | ···················· |
| W | 5472 | hCoV-19/India/MH-ACTREC-295/2020\|EPI_ISL_699947\|20 | ···················· | ···················· | ···················· | ···················· | ···················· | ···················· |
| W | 5473 | hCoV-19/India/MH-ACTREC-296/2020\|EPI_ISL_699948\|20 | ···················· | ···················· | ···················· | ···················· | ···················· | ···················· |
| W | 5474 | hCoV-19/India/MH-ACTREC-297/2020\|EPI_ISL_699949\|20 | ···················· | ···················· | ···················· | ···················· | ···················· | ···················· |
| W | 5475 | hCoV-19/India/MH-ACTREC-298/2020\|EPI_ISL_699950\|20 | ···················· | ···················· | ···················· | ···················· | ···················· | ···················· |
| W | 5476 | hCoV-19/India/MH-ACTREC-299/2020\|EPI_ISL_699951\|20 | ···················· | ···················· | ···················· | ···················· | ···················· | ···················· |
| W | 5477 | hCoV-19/India/MH-ACTREC-301/2020\|EPI_ISL_699952\|20 | ···················· | ···················· | ···················· | ···················· | ···················· | ···················· |
| W | 5478 | hCoV-19/India/MH-ACTREC-302/2020\|EPI_ISL_699953\|20 | ···················· | ···················· | ···················· | ···················· | ···················· | ···················· |
| W | 5479 | hCoV-19/India/MH-ACTREC-303/2020\|EPI_ISL_699954\|20 | ···················· | ···················· | ···················· | ···················· | ···················· | ···················· |
| W | 5480 | hCoV-19/India/MH-ACTREC-304/2020\|EPI_ISL_699955\|20 | ···················· | ···················· | ···A················ | ···················· | ···················· | ···················· |
| W | 5481 | hCoV-19/India/MH-ACTREC-305/2020\|EPI_ISL_699956\|20 | ···················· | ···················· | ···················· | ···················· | ···················· | ···················· |
| W | 5482 | hCoV-19/India/MH-ACTREC-306/2020\|EPI_ISL_699957\|20 | ···················· | ···················· | ···················· | ···················· | ···················· | ···················· |
| W | 5483 | hCoV-19/India/MH-ACTREC-307/2020\|EPI_ISL_699958\|20 | ···················· | ···················· | ···················· | ···················· | ···················· | ···················· |
| W | 5484 | hCoV-19/India/MH-ACTREC-308/2020\|EPI_ISL_699959\|20 | ···················· | ···················· | ···················· | ···················· | ···················· | ···················· |
| W | 5485 | hCoV-19/India/MH-ACTREC-309/2020\|EPI_ISL_699960\|20 | ···················· | ···················· | ···················· | ···················· | ···················· | ···················· |
| W | 5486 | hCoV-19/India/MH-ACTREC-310/2020\|EPI_ISL_699961\|20 | ···················· | ···················· | ···················· | ···················· | ···················· | ···················· |
| W | 5487 | hCoV-19/India/MH-ACTREC-313/2020\|EPI_ISL_699962\|20 | ···················· | ···················· | ···················· | ···················· | ···················· | ···················· |
| W | 5488 | hCoV-19/India/MH-ACTREC-314/2020\|EPI_ISL_699963\|20 | ···················· | ···················· | ···················· | ···················· | ···················· | ···················· |
| W | 5489 | hCoV-19/India/MH-ACTREC-315/2020\|EPI_ISL_699964\|20 | ···················· | ···················· | ···················· | ···················· | ···················· | ···················· |
| W | 5490 | hCoV-19/India/MH-ACTREC-316/2020\|EPI_ISL_699965\|20 | ···················· | ···················· | ···················· | ···················· | ···················· | ···················· |
| W | 5491 | hCoV-19/India/MH-ACTREC-317/2020\|EPI_ISL_699966\|20 | ···················· | ···················· | ···················· | ···················· | ···················· | ···················· |
| W | 5492 | hCoV-19/India/MH-ACTREC-318/2020\|EPI_ISL_699967\|20 | ···················· | ···················· | ···················· | ···················· | ···················· | ···················· |
| W | 5493 | hCoV-19/India/MH-ACTREC-319/2020\|EPI_ISL_699968\|20 | ···················· | ···················· | ···················· | ···················· | ···················· | ···················· |
| W | 5494 | hCoV-19/India/MH-ACTREC-320/2020\|EPI_ISL_699969\|20 | ···················· | ···················· | ···················· | ···················· | ···················· | ···················· |
| W | 5495 | hCoV-19/India/MH-ACTREC-321/2020\|EPI_ISL_699970\|20 | ···················· | ···················· | ···················· | ···················· | ···················· | ···················· |
| W | 5496 | hCoV-19/India/MH-ACTREC-322/2020\|EPI_ISL_699971\|20 | ···················· | ···················· | ···················· | ···················· | ···················· | ···················· |
| W | 5497 | hCoV-19/India/MH-ACTREC-323/2020\|EPI_ISL_699972\|20 | ···················· | ···················· | ···················· | ···················· | ···················· | ···················· |

| | | | | | | | | |
|---|---|---|---|---|---|---|---|---|
| W | 5498 | hCoV-19/India/MH-ACTREC-324/2020\|EPI_ISL_699973\|20 | ···················· | ···················· | ···················· | ···················· | ···················· | ···················· |
| W | 5499 | hCoV-19/India/MH-ACTREC-325/2020\|EPI_ISL_699974\|20 | ···················· | ···················· | ···················· | ···················· | ···················· | ···················· |
| W | 5500 | hCoV-19/India/MH-ACTREC-326/2020\|EPI_ISL_699975\|20 | ···················· | ···················· | ···················· | ···················· | ···················· | ···················· |
| W | 5501 | hCoV-19/India/MH-ACTREC-327/2020\|EPI_ISL_699976\|20 | ···················· | ···················· | ···················· | ···················· | ···················· | ···················· |
| W | 5502 | hCoV-19/India/MH-ACTREC-328/2020\|EPI_ISL_699977\|20 | ···················· | ···················· | ···················· | ···················· | ···················· | ···················· |
| W | 5503 | hCoV-19/India/MH-ACTREC-329/2020\|EPI_ISL_699978\|20 | ···················· | ···················· | ···················· | ···················· | ···················· | ···················· |
| W | 5504 | hCoV-19/India/MH-ACTREC-330/2020\|EPI_ISL_699979\|20 | ···················· | ···················· | ···················· | ···················· | ···················· | ···················· |
| W | 5505 | hCoV-19/India/MH-ACTREC-331/2020\|EPI_ISL_699980\|20 | ···················· | ···················· | ···················· | ···················· | ···················· | ···················· |
| W | 5506 | hCoV-19/India/MH-ACTREC-332/2020\|EPI_ISL_699981\|20 | ···················· | ···················· | ···················· | ···················· | ···················· | ···················· |
| W | 5507 | hCoV-19/India/MH-ACTREC-333/2020\|EPI_ISL_699982\|20 | ···················· | ···················· | ···················· | ···················· | ···················· | ···················· |
| W | 5508 | hCoV-19/India/MH-ACTREC-334/2020\|EPI_ISL_699983\|20 | ···················· | ···················· | ···················· | ···················· | ···················· | ···················· |
| W | 5509 | hCoV-19/India/MH-ACTREC-335/2020\|EPI_ISL_699984\|20 | ···················· | ·····A·············· | ···················· | ···················· | ···················· | ···················· |
| W | 5510 | hCoV-19/India/MH-ACTREC-336/2020\|EPI_ISL_699985\|20 | ···················· | ···················· | ···················· | ···················· | ···················· | ···················· |
| W | 5511 | hCoV-19/India/MH-ACTREC-337/2020\|EPI_ISL_699986\|20 | ···················· | ···················· | ···················· | ···················· | ···················· | ···················· |
| W | 5512 | hCoV-19/India/MH-ACTREC-338/2020\|EPI_ISL_699987\|20 | ···················· | ···················· | ···················· | ···················· | ···················· | ···················· |
| W | 5513 | hCoV-19/India/MH-ACTREC-339/2020\|EPI_ISL_699988\|20 | ···················· | ···················· | ···················· | ···················· | ···················· | ···················· |
| W | 5514 | hCoV-19/India/MH-ACTREC-340/2020\|EPI_ISL_699989\|20 | ···················· | ···················· | ···················· | ···················· | ···················· | ···················· |
| W | 5515 | hCoV-19/India/MH-ACTREC-341/2020\|EPI_ISL_699990\|20 | ···················· | ···················· | ···················· | ···················· | ···················· | ···················· |
| W | 5516 | hCoV-19/India/MH-ACTREC-342/2020\|EPI_ISL_699991\|20 | ···················· | ···················· | ···················· | ···················· | ···················· | ···················· |
| W | 5517 | hCoV-19/India/MH-ACTREC-343/2020\|EPI_ISL_699992\|20 | ···················· | ···················· | ···················· | ···················· | ···················· | ···················· |
| W | 5518 | hCoV-19/India/MH-ACTREC-344/2020\|EPI_ISL_699993\|20 | ···················· | ···················· | ···················· | ···················· | ···················· | ···················· |
| W | 5519 | hCoV-19/India/MH-ACTREC-345/2020\|EPI_ISL_699994\|20 | ···················· | ···················· | ···················· | ···················· | ···················· | ···················· |
| W | 5520 | hCoV-19/India/MH-ACTREC-346/2020\|EPI_ISL_699995\|20 | ···················· | ···················· | ···················· | ···················· | ···················· | ···················· |
| W | 5521 | hCoV-19/India/MH-ACTREC-347/2020\|EPI_ISL_699996\|20 | ···················· | ···················· | ···················· | ···················· | ···················· | ···················· |
| W | 5522 | hCoV-19/India/MH-ACTREC-348/2020\|EPI_ISL_699997\|20 | ···················· | ···················· | ···················· | ···················· | ···················· | ···················· |
| W | 5523 | hCoV-19/India/MH-ACTREC-349/2020\|EPI_ISL_699998\|20 | ···················· | ···················· | ···················· | ···················· | ···················· | ···················· |
| W | 5524 | hCoV-19/India/MH-ACTREC-350/2020\|EPI_ISL_699999\|20 | ···················· | ···················· | ···················· | ···················· | ···················· | ···················· |
| W | 5525 | hCoV-19/England/ALDP-BBAC15/2020\|EPI_ISL_701900\|20 | ···················· | ···················· | ···················· | ···················· | ···················· | ···················· |
| W | 5526 | hCoV-19/England/ALDP-BBAF85/2020\|EPI_ISL_701901\|20 | ···················· | ···················· | ···················· | ···················· | ···················· | ···················· |
| W | 5527 | hCoV-19/England/ALDP-BBA750/2020\|EPI_ISL_701902\|20 | ···················· | ···················· | ···················· | ···················· | ···················· | ···················· |
| W | 5528 | hCoV-19/England/ALDP-BBAFFE/2020\|EPI_ISL_701903\|20 | ···················· | ···················· | ···················· | ···················· | ···················· | ···················· |
| W | 5529 | hCoV-19/England/ALDP-BB4F8B/2020\|EPI_ISL_701904\|20 | ···················· | ···················· | ···················· | ···················· | ···················· | ···················· |
| W | 5530 | hCoV-19/England/ALDP-BBB33D/2020\|EPI_ISL_701905\|20 | ···················· | ·A·················· | ···················· | ···················· | ···················· | ···················· |
| W | 5531 | hCoV-19/England/ALDP-BB504B/2020\|EPI_ISL_701906\|20 | ···················· | ·A·················· | ···················· | ···················· | ···················· | ···················· |
| W | 5532 | hCoV-19/England/ALDP-BB5218/2020\|EPI_ISL_701907\|20 | ···················· | ···················· | ···················· | ···················· | ···················· | ···················· |
| W | 5533 | hCoV-19/England/ALDP-BB3CB2/2020\|EPI_ISL_701908\|20 | ···················· | ···················· | ···················· | ···················· | ···················· | ···················· |
| W | 5534 | hCoV-19/England/ALDP-BBAFC1/2020\|EPI_ISL_701909\|20 | ···················· | ···················· | ···················· | ···················· | ···················· | ···················· |
| W | 5535 | hCoV-19/England/ALDP-BBA3E3/2020\|EPI_ISL_701910\|20 | ···················· | ···················· | ···················· | ···················· | ···················· | ···················· |
| W | 5536 | hCoV-19/England/ALDP-BB3CFE/2020\|EPI_ISL_701911\|20 | ···················· | ···················· | ···················· | ···················· | ···················· | ···················· |
| W | 5537 | hCoV-19/England/ALDP-BBAC42/2020\|EPI_ISL_701934\|20 | ···················· | ···················· | ···················· | ···················· | ···················· | ···················· |
| W | 5538 | hCoV-19/England/ALDP-BB9FB3/2020\|EPI_ISL_701935\|20 | ···················· | ···················· | ···················· | ···················· | ···················· | ···················· |
| W | 5539 | hCoV-19/England/ALDP-BBB17F/2020\|EPI_ISL_701936\|20 | ···················· | ···················· | ···················· | ···················· | ···················· | ···················· |
| W | 5540 | hCoV-19/England/ALDP-BBA055/2020\|EPI_ISL_701938\|20 | ···················· | ···················· | ···················· | ···················· | ···················· | ···················· |
| W | 5541 | hCoV-19/England/ALDP-BBB063/2020\|EPI_ISL_701939\|20 | ···················· | ···················· | ···················· | ···················· | ···················· | ···················· |
| W | 5542 | hCoV-19/England/ALDP-BBB476/2020\|EPI_ISL_701940\|20 | ···················· | ···················· | ···················· | ···················· | ···················· | ···················· |
| W | 5543 | hCoV-19/England/ALDP-BB9FFF/2020\|EPI_ISL_701942\|20 | ···················· | ···················· | ···················· | ···················· | ···················· | ···················· |
| W | 5544 | hCoV-19/England/ALDP-BB3AE5/2020\|EPI_ISL_701943\|20 | ···················· | ···················· | ···················· | ···················· | ·····T·············· | ···················· |
| W | 5545 | hCoV-19/England/ALDP-BB3C85/2020\|EPI_ISL_701944\|20 | ···················· | ···················· | ···················· | ···················· | ···················· | ···················· |
| W | 5546 | hCoV-19/England/ALDP-BB512A/2020\|EPI_ISL_701945\|20 | ···················· | ···················· | ···················· | ···················· | ···················· | ···················· |
| W | 5547 | hCoV-19/England/ALDP-BBA1BC/2020\|EPI_ISL_701946\|20 | ···················· | ···················· | ···················· | ···················· | ···················· | ···················· |
| W | 5548 | hCoV-19/England/ALDP-BB547C/2020\|EPI_ISL_701948\|20 | ···················· | ···················· | ···················· | ···················· | ···················· | ···················· |
| W | 5549 | hCoV-19/England/ALDP-BBAF3A/2020\|EPI_ISL_701950\|20 | ···················· | ···················· | ···················· | ···················· | ···················· | ···················· |
| W | 5550 | hCoV-19/England/QEUH-BC35F0/2020\|EPI_ISL_704110\|20 | ···················· | ···················· | ···················· | ···················· | ···················· | ···················· |
| W | 5551 | hCoV-19/England/QEUH-BC4BC0/2020\|EPI_ISL_704114\|20 | ···················· | ···················· | ···················· | ···················· | ···················· | ···················· |
| W | 5552 | hCoV-19/England/QEUH-BC3BC1/2020\|EPI_ISL_704116\|20 | ···················· | ···················· | ···················· | ···················· | ···················· | ···················· |
| W | 5553 | hCoV-19/England/QEUH-BCB886/2020\|EPI_ISL_704117\|20 | ···················· | ···················· | ···················· | ···················· | ···················· | ···················· |
| W | 5554 | hCoV-19/England/QEUH-BC3A2E/2020\|EPI_ISL_704121\|20 | ···················· | ···················· | ···················· | ···················· | ···················· | ···················· |
| W | 5555 | hCoV-19/England/QEUH-BC3C19/2020\|EPI_ISL_704122\|20 | ···················· | ···················· | ···················· | ···················· | ···················· | ···················· |
| W | 5556 | hCoV-19/England/QEUH-BC3CFB/2020\|EPI_ISL_704124\|20 | ···················· | ···················· | ···················· | ···················· | ···················· | ···················· |
| W | 5557 | hCoV-19/England/QEUH-BCC700/2020\|EPI_ISL_704125\|20 | ···················· | ···················· | ···················· | ···················· | ···················· | ···················· |
| W | 5558 | hCoV-19/England/QEUH-BCB938/2020\|EPI_ISL_704127\|20 | ···················· | ···················· | ···················· | ···················· | ···················· | ···················· |
| W | 5559 | hCoV-19/England/QEUH-BCBCC6/2020\|EPI_ISL_704128\|20 | ···················· | ···················· | ···················· | ···················· | ···················· | ···················· |
| W | 5560 | hCoV-19/England/QEUH-BC3DF8/2020\|EPI_ISL_704138\|20 | ···················· | ···················· | ···················· | ···················· | ···················· | ···················· |
| W | 5561 | hCoV-19/England/QEUH-BCC13F/2020\|EPI_ISL_704140\|20 | ···················· | ···················· | ···················· | ···················· | ···················· | ···················· |
| W | 5562 | hCoV-19/England/QEUH-BC3684/2020\|EPI_ISL_704143\|20 | ···················· | ···················· | ···················· | ···················· | ···················· | ···················· |
| W | 5563 | hCoV-19/England/QEUH-BCB640/2020\|EPI_ISL_704144\|20 | ···················· | ···················· | ···················· | ···················· | ···················· | ···················· |
| W | 5564 | hCoV-19/England/QEUH-BCB68C/2020\|EPI_ISL_704146\|20 | ···················· | ···················· | ···················· | ···················· | ···················· | ···················· |
| W | 5565 | hCoV-19/England/QEUH-BC3639/2020\|EPI_ISL_704151\|20 | ···················· | ···················· | ···················· | ···················· | ···················· | ···················· |
| W | 5566 | hCoV-19/England/QEUH-BCC014/2020\|EPI_ISL_704152\|20 | ···················· | ···················· | ···················· | ···················· | ···················· | ···················· |
| W | 5567 | hCoV-19/England/QEUH-BC3CEC/2020\|EPI_ISL_704154\|20 | ···················· | ···················· | ···················· | ···················· | ···················· | ···················· |
| W | 5568 | hCoV-19/England/QEUH-BCC0D8/2020\|EPI_ISL_704157\|20 | ···················· | ···················· | ···················· | ···················· | ···················· | ···················· |
| W | 5569 | hCoV-19/England/QEUH-BC3B67/2020\|EPI_ISL_704159\|20 | ···················· | ·A·················· | ···················· | ···················· | ···················· | ···················· |
| W | 5570 | hCoV-19/England/QEUH-BCB437/2020\|EPI_ISL_704160\|20 | ···················· | ···················· | ···················· | ···················· | ···················· | ···················· |
| W | 5571 | hCoV-19/England/QEUH-BC3727/2020\|EPI_ISL_704162\|20 | ···················· | ···················· | ···················· | ···················· | ···················· | ···················· |
| W | 5572 | hCoV-19/England/QEUH-BC3D16/2020\|EPI_ISL_704164\|20 | ···················· | ···················· | ···················· | ···················· | ···················· | ···················· |
| W | 5573 | hCoV-19/England/QEUH-BC3666/2020\|EPI_ISL_704165\|20 | ···················· | ···················· | ···················· | ···················· | ···················· | ···················· |
| W | 5574 | hCoV-19/England/QEUH-BCC050/2020\|EPI_ISL_704167\|20 | ···················· | ···················· | ···················· | ···················· | ···················· | ···················· |
| W | 5575 | hCoV-19/England/QEUH-BCC08D/2020\|EPI_ISL_704169\|20 | ···················· | ···················· | ···················· | ···················· | ·····T·············· | ···················· |
| W | 5576 | hCoV-19/England/QEUH-BCBE0C/2020\|EPI_ISL_704170\|20 | ···················· | ···················· | ···················· | ···················· | ···················· | ···················· |
| W | 5577 | hCoV-19/England/QEUH-BCBFAF/2020\|EPI_ISL_704172\|20 | ···················· | ···················· | ···················· | ···················· | ···················· | ···················· |

| | | | | | | | |
|---|---|---|---|---|---|---|---|
| W | 5578 hCoV-19/England/QEUH-BCB491/2020\|EPI_ISL_704173\|20 | ···················· | ···················· | ···················· | ···················· | ···················· | ···················· |
| W | 5579 hCoV-19/England/QEUH-BC3E7D/2020\|EPI_ISL_704177\|20 | ···················· | ···················· | ···················· | ···················· | ···················· | ···················· |
| W | 5580 hCoV-19/England/QEUH-BC3DBC/2020\|EPI_ISL_704178\|20 | ···················· | ···················· | ···················· | ···················· | ···················· | ···················· |
| W | 5581 hCoV-19/England/QEUH-BC4BEE/2020\|EPI_ISL_704180\|20 | ···················· | ···················· | ···················· | ···················· | ···················· | ···················· |
| W | 5582 hCoV-19/England/QEUH-BC3C55/2020\|EPI_ISL_704182\|20 | ···················· | ···················· | ···················· | ···················· | ···················· | ···················· |
| W | 5583 hCoV-19/England/QEUH-BC362A/2020\|EPI_ISL_704183\|20 | ···················· | ···················· | ···················· | ···················· | ···················· | ···················· |
| W | 5584 hCoV-19/England/QEUH-BCB4CE/2020\|EPI_ISL_704185\|20 | ···················· | ···················· | ···················· | ···················· | ···················· | ···················· |
| W | 5585 hCoV-19/England/QEUH-BCC72E/2020\|EPI_ISL_704186\|20 | ···················· | ···················· | ···················· | ···················· | ···················· | ···················· |
| W | 5586 hCoV-19/England/QEUH-BCB464/2020\|EPI_ISL_704188\|20 | ···················· | ···················· | ···················· | ···················· | ···················· | ···················· |
| W | 5587 hCoV-19/England/QEUH-BC4AC3/2020\|EPI_ISL_704189\|20 | ···················· | ···················· | ···················· | ···················· | ···················· | ···················· |
| W | 5588 hCoV-19/Wales/PHWC-492166/2020\|EPI_ISL_705440\|2020 | ···················· | ···················· | ···················· | ···················· | ···················· | ···················· |
| W | 5590 hCoV-19/Wales/PHWC-492333/2020\|EPI_ISL_705442\|2020 | ···················· | ···················· | ···················· | ···················· | ···················· | ···················· |
| W | 5591 hCoV-19/England/QEUH-A35F1A/2020\|EPI_ISL_705443\|20 | ···················· | ···················· | ···················· | ···················· | ···················· | ···················· |
| W | 5592 hCoV-19/Wales/QEUH-A62020/2020\|EPI_ISL_705444\|2020 | ···················· | ···················· | ···················· | ···················· | ···················· | ···················· |
| W | 5593 hCoV-19/England/LIVE-DC2823/2020\|EPI_ISL_705471\|20 | ···················· | ···················· | ···················· | ···················· | ···················· | ···················· |
| W | 5594 hCoV-19/England/LOND-1251AB2/2020\|EPI_ISL_705472\|2 | ···················· | ·A·················· | ···················· | ···················· | ···················· | ··A················· |
| W | 5595 hCoV-19/England/MILK-A91671/2020\|EPI_ISL_705473\|20 | ···················· | ·A·················· | ···················· | ···················· | ···················· | ··A················· |
| W | 5596 hCoV-19/England/MILK-B3A785/2020\|EPI_ISL_705474\|20 | ···················· | ·A·················· | ···················· | ···················· | ···················· | ··A················· |
| W | 5597 hCoV-19/England/MILK-B3A7A3/2020\|EPI_ISL_705475\|20 | ···················· | ·A·················· | ···················· | ···················· | ···················· | ··A················· |
| W | 5598 hCoV-19/England/MILK-B3B21A/2020\|EPI_ISL_705476\|20 | ···················· | ·A·················· | ···················· | ···················· | ···················· | ··A················· |
| W | 5599 hCoV-19/England/MILK-B3B256/2020\|EPI_ISL_705477\|20 | ···················· | ·A·················· | ···················· | ···················· | ···················· | ··A················· |
| W | 5600 hCoV-19/England/MILK-B3B274/2020\|EPI_ISL_705478\|20 | ···················· | ·A·················· | ···················· | ···················· | ···················· | ··A················· |
| W | 5601 hCoV-19/England/MILK-B3C01F/2020\|EPI_ISL_705479\|20 | ···················· | ·A·················· | ···················· | ···················· | ···················· | ··A················· |
| W | 5602 hCoV-19/England/MILK-B3C03D/2020\|EPI_ISL_705480\|20 | ···················· | ·A·················· | ···················· | ···················· | ···················· | ··A················· |
| W | 5603 hCoV-19/England/MILK-B3C7CF/2020\|EPI_ISL_705481\|20 | ···················· | ·A·················· | ···················· | ···················· | ···················· | ··A················· |
| W | 5604 hCoV-19/England/MILK-B3CA3F/2020\|EPI_ISL_705482\|20 | ···················· | ·A·················· | ···················· | ···················· | ···················· | ··A················· |
| W | 5605 hCoV-19/England/MILK-B3CDBE/2020\|EPI_ISL_705483\|20 | ···················· | ·A·················· | ···················· | ···················· | ···················· | ··A················· |
| W | 5606 hCoV-19/England/MILK-B3DC74/2020\|EPI_ISL_705484\|20 | ···················· | ·A·················· | ···················· | ···················· | ···················· | ··A················· |
| W | 5607 hCoV-19/England/MILK-B3DCED/2020\|EPI_ISL_705485\|20 | ···················· | ·A·················· | ···················· | ···················· | ···················· | ··A················· |
| W | 5608 hCoV-19/England/NORT-1B71CAE/2020\|EPI_ISL_705486\|2 | ···················· | ···················· | ···················· | ···················· | ···················· | ···················· |
| W | 5609 hCoV-19/England/NOTT-11FAA5/2020\|EPI_ISL_705487\|20 | ···················· | ·A·················· | ···················· | ···················· | ···················· | ··A················· |
| W | 5610 hCoV-19/Wales/PHWC-490353/2020\|EPI_ISL_705488\|2020 | ···················· | ···················· | ···················· | ···················· | ···················· | ···················· |
| W | 5611 hCoV-19/England/QEUH-A6205D/2020\|EPI_ISL_705489\|20 | ···················· | ·A·················· | ···················· | ···················· | ···················· | ··A················· |
| W | 5612 hCoV-19/Wales/PHWC-491D09/2020\|EPI_ISL_706770\|2020 | ···················· | ···················· | ···················· | ···················· | ···················· | ···················· |
| W | 5613 hCoV-19/Wales/PHWC-491D27/2020\|EPI_ISL_706771\|2020 | ···················· | ···················· | ···················· | ···················· | ···················· | ···················· |
| W | 5614 hCoV-19/Wales/PHWC-491D36/2020\|EPI_ISL_706772\|2020 | ···················· | ···················· | ···················· | ···················· | ···················· | ···················· |
| W | 5615 hCoV-19/Wales/PHWC-491D45/2020\|EPI_ISL_706773\|2020 | ···················· | ···················· | ···················· | ···················· | ···················· | ···················· |
| W | 5617 hCoV-19/Wales/PHWC-491D63/2020\|EPI_ISL_706775\|2020 | ···················· | ···················· | ···················· | ···················· | ···················· | ···················· |
| W | 5618 hCoV-19/Wales/PHWC-491D81/2020\|EPI_ISL_706776\|2020 | ···················· | ···················· | ···················· | ···················· | ···················· | ···················· |
| W | 5619 hCoV-19/Wales/PHWC-491D90/2020\|EPI_ISL_706777\|2020 | ···················· | ···················· | ···················· | ···················· | ···················· | NN·················· |
| W | 5621 hCoV-19/Wales/PHWC-491DBE/2020\|EPI_ISL_706779\|2020 | ···················· | ···················· | ···················· | ···················· | ···················· | ···················· |
| W | 5624 hCoV-19/Wales/PHWC-491E06/2020\|EPI_ISL_706782\|2020 | ···················· | ···················· | ···················· | ···················· | ···················· | ···················· |
| W | 5628 hCoV-19/Wales/PHWC-491E42/2020\|EPI_ISL_706786\|2020 | ···················· | ···················· | ···················· | ···················· | ···················· | ···················· |
| W | 5629 hCoV-19/Wales/PHWC-491E51/2020\|EPI_ISL_706787\|2020 | ···················· | ···················· | ···················· | ···················· | ···················· | ···················· |
| W | 5631 hCoV-19/Wales/PHWC-491E8E/2020\|EPI_ISL_706789\|2020 | ···················· | ···················· | ···················· | ···················· | ···················· | ···················· |
| W | 5633 hCoV-19/United Arab Emirates/0382/2020\|EPI_ISL_698 | ···················· | ···················· | ···················· | ···················· | ···················· | ···················· |
| W | 5634 hCoV-19/United Arab Emirates/0383/2020\|EPI_ISL_698 | ···················· | ···················· | ···················· | ···················· | ···················· | ···················· |
| W | 5635 hCoV-19/United Arab Emirates/0384/2020\|EPI_ISL_698 | ···················· | ···················· | ···················· | ···················· | ···················· | ···················· |
| W | 5636 hCoV-19/United Arab Emirates/0385/2020\|EPI_ISL_698 | ···················· | ···················· | ···················· | ···················· | ···················· | ···················· |
| W | 5637 hCoV-19/United Arab Emirates/0386/2020\|EPI_ISL_698 | ···················· | ···················· | ···················· | ···················· | ···················· | ···················· |
| W | 5638 hCoV-19/United Arab Emirates/0387/2020\|EPI_ISL_698 | ···················· | ···················· | ···················· | ···················· | ···················· | ···················· |
| W | 5639 hCoV-19/United Arab Emirates/0388/2020\|EPI_ISL_698 | ···················· | ···················· | ···················· | ···················· | ···················· | ···················· |
| W | 5640 hCoV-19/United Arab Emirates/0389/2020\|EPI_ISL_698 | ···················· | ···················· | ···················· | ···················· | ···················· | ···················· |
| W | 5641 hCoV-19/United Arab Emirates/0390/2020\|EPI_ISL_698 | ···················· | ···················· | ···················· | ···················· | ···················· | ···················· |
| W | 5642 hCoV-19/United Arab Emirates/0391/2020\|EPI_ISL_698 | ···················· | ···················· | ···················· | ···················· | ···················· | ···················· |
| W | 5643 hCoV-19/United Arab Emirates/0392/2020\|EPI_ISL_698 | ···················· | ···················· | ···················· | ···················· | ···················· | ···················· |
| W | 5644 hCoV-19/United Arab Emirates/0393/2020\|EPI_ISL_698 | ···················· | ···················· | ···················· | ···················· | ···················· | ···················· |
| W | 5645 hCoV-19/United Arab Emirates/0394/2020\|EPI_ISL_698 | ···················· | ···················· | ···················· | ···················· | ···················· | ···················· |
| W | 5646 hCoV-19/United Arab Emirates/0395/2020\|EPI_ISL_698 | ···················· | ···················· | ···················· | ···················· | ···················· | ···················· |
| W | 5647 hCoV-19/United Arab Emirates/0396/2020\|EPI_ISL_698 | ···················· | ···················· | ···················· | ···················· | ···················· | ···················· |
| W | 5648 hCoV-19/United Arab Emirates/0397/2020\|EPI_ISL_698 | ···················· | ······A············· | ···················· | ···················· | ···················· | ···················· |
| W | 5649 hCoV-19/United Arab Emirates/0398/2020\|EPI_ISL_698 | ···················· | ···················· | ···················· | ···················· | ···················· | ···················· |
| W | 5650 hCoV-19/United Arab Emirates/0399/2020\|EPI_ISL_698 | ···················· | ···················· | ···················· | ···················· | ···················· | ···················· |
| W | 5651 hCoV-19/United Arab Emirates/0400/2020\|EPI_ISL_698 | ···················· | ···················· | ···················· | ···················· | ···················· | ···················· |
| W | 5652 hCoV-19/United Arab Emirates/0401/2020\|EPI_ISL_698 | ···················· | ···················· | ···················· | ···················· | ···················· | ···················· |
| W | 5653 hCoV-19/United Arab Emirates/0402/2020\|EPI_ISL_698 | ···················· | ···················· | ···················· | ···················· | ···················· | ···················· |
| W | 5654 hCoV-19/United Arab Emirates/0403/2020\|EPI_ISL_698 | ···················· | ···················· | ···················· | ···················· | ···················· | ···················· |
| W | 5655 hCoV-19/India/MH-ACTREC-235/2020\|EPI_ISL_699888\|20 | ···················· | ···················· | ···················· | ···················· | ···················· | ···················· |
| W | 5656 hCoV-19/India/MH-ACTREC-236/2020\|EPI_ISL_699889\|20 | ···················· | ···················· | ···················· | ···················· | ···················· | ···················· |
| W | 5657 hCoV-19/India/MH-ACTREC-237/2020\|EPI_ISL_699890\|20 | ···················· | ···················· | ···················· | ···················· | ···················· | ···················· |
| W | 5658 hCoV-19/India/MH-ACTREC-238/2020\|EPI_ISL_699891\|20 | ···················· | ···················· | ···················· | ···················· | ···················· | ···················· |
| W | 5659 hCoV-19/India/MH-ACTREC-239/2020\|EPI_ISL_699892\|20 | ···················· | ···················· | ···················· | ···················· | ···················· | ···················· |
| W | 5660 hCoV-19/India/MH-ACTREC-240/2020\|EPI_ISL_699893\|20 | ···················· | ···················· | ···················· | ···················· | ···················· | ···················· |
| W | 5661 hCoV-19/India/MH-ACTREC-241/2020\|EPI_ISL_699894\|20 | ···················· | ···················· | ···················· | ···················· | ···················· | ···················· |
| W | 5662 hCoV-19/India/MH-ACTREC-242/2020\|EPI_ISL_699895\|20 | ···················· | ···················· | ···················· | ···················· | ···················· | ···················· |
| W | 5663 hCoV-19/India/MH-ACTREC-243/2020\|EPI_ISL_699896\|20 | ···················· | ···················· | ···················· | ···················· | ···················· | ···················· |
| W | 5664 hCoV-19/India/MH-ACTREC-244/2020\|EPI_ISL_699897\|20 | ···················· | ···················· | ···················· | ···················· | ···················· | ···················· |
| W | 5665 hCoV-19/India/MH-ACTREC-245/2020\|EPI_ISL_699898\|20 | ···················· | ···················· | ···················· | ···················· | ···················· | ···················· |
| W | 5666 hCoV-19/India/MH-ACTREC-246/2020\|EPI_ISL_699899\|20 | ···················· | ···················· | ···················· | ···················· | ···················· | ···················· |
| W | 5667 hCoV-19/India/MH-ACTREC-247/2020\|EPI_ISL_699900\|20 | ···················· | ···················· | ···················· | ···················· | ···················· | ···················· |

| | | | | | | | | |
|---|---|---|---|---|---|---|---|---|
| W | 5668 | hCoV-19/India/MH-ACTREC-249/2020\|EPI_ISL_699901\|20 | .................... | .................... | .................... | .................... | .................... | .................... |
| W | 5669 | hCoV-19/India/MH-ACTREC-250/2020\|EPI_ISL_699902\|20 | .................... | .................... | .................... | .................... | .................... | .................... |
| W | 5670 | hCoV-19/India/MH-ACTREC-251/2020\|EPI_ISL_699903\|20 | .................... | .................... | .................... | .................... | .................... | .................... |
| W | 5672 | hCoV-19/India/MH-ACTREC-254/2020\|EPI_ISL_699906\|20 | .................... | .................... | ...T................ | .................... | .................... | .................... |
| W | 5673 | hCoV-19/India/MH-ACTREC-255/2020\|EPI_ISL_699907\|20 | .................... | .................... | .................... | .................... | .................... | .................... |
| W | 5674 | hCoV-19/India/MH-ACTREC-256/2020\|EPI_ISL_699908\|20 | .................... | .................... | .................... | .................... | .................... | .................... |
| W | 5675 | hCoV-19/India/MH-ACTREC-257/2020\|EPI_ISL_699909\|20 | .................... | .................... | .................... | .................... | .................... | .................... |
| W | 5676 | hCoV-19/India/MH-ACTREC-258/2020\|EPI_ISL_699910\|20 | .................... | .................... | .................... | .................... | .................... | .................... |
| W | 5677 | hCoV-19/India/MH-ACTREC-259/2020\|EPI_ISL_699911\|20 | .................... | .................... | .................... | .................... | .................... | .................... |
| W | 5678 | hCoV-19/India/MH-ACTREC-260/2020\|EPI_ISL_699912\|20 | .................... | .................... | .................... | .................... | .................... | .................... |
| W | 5679 | hCoV-19/India/MH-ACTREC-261/2020\|EPI_ISL_699913\|20 | .................... | .................... | .................... | .................... | .................... | .................... |
| W | 5680 | hCoV-19/India/MH-ACTREC-262/2020\|EPI_ISL_699914\|20 | .................... | .................... | .................... | .................... | .................... | .................... |
| W | 5681 | hCoV-19/India/MH-ACTREC-263/2020\|EPI_ISL_699915\|20 | .................... | .................... | .................... | .................... | .................... | .................... |
| W | 5682 | hCoV-19/India/MH-ACTREC-264/2020\|EPI_ISL_699916\|20 | .................... | .................... | .................... | .................... | .................... | .................... |
| W | 5684 | hCoV-19/India/MH-ACTREC-266/2020\|EPI_ISL_699918\|20 | .................... | .................... | .................... | .................... | .................... | .................... |
| W | 5685 | hCoV-19/India/MH-ACTREC-267/2020\|EPI_ISL_699919\|20 | .................... | .................... | .................... | .................... | .................... | .................... |
| W | 5686 | hCoV-19/India/MH-ACTREC-268/2020\|EPI_ISL_699920\|20 | .................... | .................... | .................... | .................... | .................... | .................... |
| W | 5688 | hCoV-19/England/204622419/2020\|EPI_ISL_700654\|2020 | .................... | .................... | .................... | .................... | .................... | .................... |
| W | 5689 | hCoV-19/England/204620552/2020\|EPI_ISL_700671\|2020 | .................... | .................... | .................... | .................... | .................... | .................... |
| W | 5690 | hCoV-19/England/204590738/2020\|EPI_ISL_700672\|2020 | .................... | .................... | .................... | .................... | .................... | .................... |
| W | 5691 | hCoV-19/England/204590764/2020\|EPI_ISL_700673\|2020 | .................... | .................... | .................... | .................... | .................... | .................... |
| W | 5692 | hCoV-19/England/204591406/2020\|EPI_ISL_700674\|2020 | .................... | .................... | .................... | .................... | ......K............. | .................... |
| W | 5693 | hCoV-19/England/204591532/2020\|EPI_ISL_700675\|2020 | .................... | .................... | .................... | .................... | .................... | .................... |
| W | 5694 | hCoV-19/England/204591539/2020\|EPI_ISL_700676\|2020 | .................... | .................... | .................... | .................... | ......T............. | .................... |
| W | 5695 | hCoV-19/England/204591540/2020\|EPI_ISL_700677\|2020 | .................... | .................... | .................... | .................... | .................... | .................... |
| W | 5696 | hCoV-19/England/204591733/2020\|EPI_ISL_700678\|2020 | .................... | .................... | .................... | .................... | .................... | .................... |
| W | 5697 | hCoV-19/England/204600303/2020\|EPI_ISL_700679\|2020 | .................... | .................... | .................... | .................... | .................... | .................... |
| W | 5698 | hCoV-19/England/204600306/2020\|EPI_ISL_700680\|2020 | .................... | .................... | .................... | .................... | .................... | .................... |
| W | 5699 | hCoV-19/England/204600613/2020\|EPI_ISL_700681\|2020 | .................... | .................... | .................... | .................... | .................... | .................... |
| W | 5700 | hCoV-19/England/204620559/2020\|EPI_ISL_700682\|2020 | .................... | .................... | .................... | .................... | .................... | .................... |
| W | 5701 | hCoV-19/England/204620607/2020\|EPI_ISL_700683\|2020 | .................... | .................... | .................... | .................... | .................... | .................... |
| W | 5702 | hCoV-19/England/204641973/2020\|EPI_ISL_700684\|2020 | .................... | .................... | .................... | .................... | .................... | .................... |
| W | 5703 | hCoV-19/England/204642475/2020\|EPI_ISL_700685\|2020 | .................... | .................... | .................... | .................... | .................... | .................... |
| W | 5704 | hCoV-19/England/204961121/2020\|EPI_ISL_700686\|2020 | .................... | .................... | .................... | .................... | .................... | .................... |
| W | 5705 | hCoV-19/England/204961122/2020\|EPI_ISL_700687\|2020 | .................... | .................... | .................... | .................... | .................... | .................... |
| W | 5706 | hCoV-19/England/204961123/2020\|EPI_ISL_700688\|2020 | .................... | .................... | .................... | .................... | .................... | .................... |
| W | 5707 | hCoV-19/England/204961124/2020\|EPI_ISL_700689\|2020 | .................... | .................... | .................... | .................... | .................... | .................... |
| W | 5708 | hCoV-19/England/204961128/2020\|EPI_ISL_700690\|2020 | .................... | .................... | .................... | .................... | .................... | .................... |
| W | 5709 | hCoV-19/England/204961136/2020\|EPI_ISL_700691\|2020 | .................... | .................... | .................... | .................... | .................... | .................... |
| W | 5710 | hCoV-19/England/204961137/2020\|EPI_ISL_700692\|2020 | .................... | .................... | .................... | .................... | .................... | .................... |
| W | 5711 | hCoV-19/England/204961141/2020\|EPI_ISL_700693\|2020 | .................... | .................... | .................... | .................... | .................... | .................... |
| W | 5712 | hCoV-19/England/204961142/2020\|EPI_ISL_700694\|2020 | .................... | .................... | .................... | .................... | .................... | .................... |
| W | 5713 | hCoV-19/England/204961143/2020\|EPI_ISL_700695\|2020 | .................... | .................... | .................... | .................... | .................... | .................... |
| W | 5714 | hCoV-19/England/204961146/2020\|EPI_ISL_700696\|2020 | .................... | .................... | .................... | .................... | .................... | .................... |
| W | 5715 | hCoV-19/England/204961149/2020\|EPI_ISL_700697\|2020 | .................... | .................... | .................... | .................... | .................... | .................... |
| W | 5716 | hCoV-19/England/204961150/2020\|EPI_ISL_700698\|2020 | .................... | .................... | .................... | .................... | .................... | .................... |
| W | 5717 | hCoV-19/England/ALDP-BB3AD6/2020\|EPI_ISL_701913\|20 | .................... | .................... | .................... | .................... | .................... | .................... |
| W | 5718 | hCoV-19/England/ALDP-BB3C94/2020\|EPI_ISL_701914\|20 | .................... | .................... | .................... | .................... | .................... | .................... |
| W | 5719 | hCoV-19/England/ALDP-BBB32E/2020\|EPI_ISL_701915\|20 | .................... | .................... | .................... | .................... | .................... | .................... |
| W | 5720 | hCoV-19/England/ALDP-BBAF0D/2020\|EPI_ISL_701916\|20 | .................... | .................... | .................... | .................... | .................... | .................... |
| W | 5721 | hCoV-19/England/ALDP-BB3942/2020\|EPI_ISL_701917\|20 | .................... | .................... | .................... | .................... | .................... | .................... |
| W | 5722 | hCoV-19/England/ALDP-BB50D2/2020\|EPI_ISL_701918\|20 | .................... | .................... | .................... | .................... | .................... | .................... |
| W | 5723 | hCoV-19/England/ALDP-BBAC60/2020\|EPI_ISL_701919\|20 | .................... | .................... | .................... | .................... | .................... | .................... |
| W | 5724 | hCoV-19/England/ALDP-BB3B00/2020\|EPI_ISL_701920\|20 | .................... | .................... | .................... | .................... | .................... | .................... |
| W | 5725 | hCoV-19/England/ALDP-BBAF94/2020\|EPI_ISL_701922\|20 | .................... | .................... | .................... | .................... | .................... | .................... |
| W | 5726 | hCoV-19/England/ALDP-BB3A5E/2020\|EPI_ISL_701923\|20 | .................... | .................... | .................... | .................... | ......T............. | .................... |
| W | 5727 | hCoV-19/England/ALDP-BBB458/2020\|EPI_ISL_701924\|20 | .................... | .................... | .................... | .................... | .................... | .................... |
| W | 5728 | hCoV-19/England/ALDP-BB5290/2020\|EPI_ISL_701925\|20 | .................... | .................... | .................... | .................... | .................... | .................... |
| W | 5729 | hCoV-19/England/ALDP-BBA79C/2020\|EPI_ISL_701927\|20 | .................... | .................... | .................... | .................... | .................... | .................... |
| W | 5730 | hCoV-19/England/ALDP-BB3DA0/2020\|EPI_ISL_701928\|20 | .................... | .................... | .................... | .................... | .................... | .................... |
| W | 5731 | hCoV-19/England/ALDP-BBA037/2020\|EPI_ISL_701931\|20 | .................... | .................... | .................... | .................... | .................... | .................... |
| W | 5732 | hCoV-19/England/ALDP-BB9F2C/2020\|EPI_ISL_701933\|20 | .................... | .................... | .................... | .................... | .................... | .................... |
| W | 5733 | hCoV-19/England/ALDP-BBA44A/2020\|EPI_ISL_701952\|20 | .................... | .................... | .................... | .................... | .................... | .................... |
| W | 5734 | hCoV-19/England/ALDP-BB5157/2020\|EPI_ISL_701953\|20 | .................... | .................... | .................... | .................... | .................... | .................... |
| W | 5735 | hCoV-19/England/ALDP-BB5148/2020\|EPI_ISL_701954\|20 | .................... | .................... | .................... | .................... | .................... | .................... |
| W | 5736 | hCoV-19/England/ALDP-BB5281/2020\|EPI_ISL_701955\|20 | .................... | .................... | .................... | .................... | .................... | .................... |
| W | 5737 | hCoV-19/England/ALDP-BBA0FB/2020\|EPI_ISL_701956\|20 | .................... | .................... | .................... | .................... | .................... | .................... |
| W | 5738 | hCoV-19/England/ALDP-BBB045/2020\|EPI_ISL_701957\|20 | .................... | .................... | .................... | .................... | .................... | .................... |
| W | 5739 | hCoV-19/England/ALDP-BB500F/2020\|EPI_ISL_701958\|20 | .................... | .................... | .................... | .................... | .................... | .................... |
| W | 5740 | hCoV-19/England/ALDP-BB9DF5/2020\|EPI_ISL_701960\|20 | .................... | .................... | .................... | .................... | .................... | .................... |
| W | 5741 | hCoV-19/England/ALDP-BB4FD6/2020\|EPI_ISL_701962\|20 | .................... | .................... | .................... | .................... | .................... | .................... |
| W | 5742 | hCoV-19/England/ALDP-BB5184/2020\|EPI_ISL_701965\|20 | .................... | .................... | .................... | .................... | .................... | .................... |
| W | 5743 | hCoV-19/England/ALDP-BBAF76/2020\|EPI_ISL_701966\|20 | .................... | .................... | .................... | .................... | .................... | .................... |
| W | 5744 | hCoV-19/England/ALDP-BBA8E4/2020\|EPI_ISL_701967\|20 | .................... | .................... | .................... | .................... | .................... | .................... |
| W | 5745 | hCoV-19/England/ALDP-BBB142/2020\|EPI_ISL_701968\|20 | .................... | .................... | .................... | .................... | .................... | .................... |
| W | 5746 | hCoV-19/England/ALDP-BBB4B2/2020\|EPI_ISL_701970\|20 | .................... | .................... | .................... | .................... | .................... | .................... |
| W | 5747 | hCoV-19/England/ALDP-BB9F77/2020\|EPI_ISL_701971\|20 | .................... | .................... | .................... | .................... | .................... | .................... |
| W | 5748 | hCoV-19/England/ALDP-BB51DF/2020\|EPI_ISL_701972\|20 | .................... | .................... | .................... | .................... | .................... | .................... |
| W | 5749 | hCoV-19/England/ALDP-BB9ED4/2020\|EPI_ISL_701974\|20 | .................... | .................... | .................... | .................... | .................... | .................... |
| W | 5750 | hCoV-19/England/ALDP-BB35A8/2020\|EPI_ISL_701975\|20 | .................... | .................... | .................... | .................... | .................... | .................... |

W 5751 hCoV-19/England/ALDP-BB3BD3/2020|EPI_ISL_701976|20 ···················· ···················· ····················· ···················· ···················· ····················
W 5752 hCoV-19/England/ALDP-BB3CA3/2020|EPI_ISL_701978|20 ···················· ···················· ····················· ···················· ···················· ····················
W 5753 hCoV-19/England/ALDP-BB53CA/2020|EPI_ISL_701979|20 ···················· ·A·················· ····················· ···················· ···················· ···A················
W 5754 hCoV-19/England/ALDP-BB3C67/2020|EPI_ISL_701981|20 ···················· ···················· ····················· ···················· ···················· ····················
W 5755 hCoV-19/England/ALDP-BB9E98/2020|EPI_ISL_701982|20 ···················· ···················· ····················· ···················· ···················· ····················
W 5756 hCoV-19/England/ALDP-BB4F6D/2020|EPI_ISL_701983|20 ··················T· ···················· ····················· ···················· ···················· ···················· 1
W 5757 hCoV-19/England/ALDP-BB505A/2020|EPI_ISL_701984|20 ···················· ···················· ····················· ···················· ···················· ····················
W 5758 hCoV-19/England/ALDP-BB511B/2020|EPI_ISL_701985|20 ···················· ·········T·········· ····················· ···················· ···················· ····················
W 5759 hCoV-19/England/ALDP-BBB212/2020|EPI_ISL_701986|20 ···················· ···················· ····················· ···················· ···················· ····················
W 5760 hCoV-19/England/ALDP-BB37B1/2020|EPI_ISL_701987|20 ···················· ·A·················· ····················· ···················· ···················· ···A················
W 5761 hCoV-19/England/ALDP-BB3D55/2020|EPI_ISL_701988|20 ···················· ···················· ····················· ···················· ···················· ····················
W 5762 hCoV-19/England/ALDP-BBA899/2020|EPI_ISL_701989|20 ···················· ···················· ····················· ···················· ···················· ····················
W 5763 hCoV-19/England/ALDP-BBA3F2/2020|EPI_ISL_701990|20 ···················· ···················· ····················· ···················· ···················· ····················
W 5764 hCoV-19/England/ALDP-BB50B4/2020|EPI_ISL_701991|20 ···················· ···················· ····················· ···················· ···················· ····················
W 5765 hCoV-19/England/ALDP-BB4FA9/2020|EPI_ISL_701992|20 ···················· ···················· ····················· ···················· ···················· ····················
W 5766 hCoV-19/England/ALDP-BB3BE2/2020|EPI_ISL_701994|20 ···················· ···················· ····················· ···················· ···················· ····················
W 5767 hCoV-19/England/ALDP-BBB0AF/2020|EPI_ISL_701995|20 ···················· ···················· ····················· ···················· ···················· ····················
W 5768 hCoV-19/England/ALDP-BBB2F4/2020|EPI_ISL_701996|20 ···················· ·A·················· ····················· ···················· ···················· ····················
W 5769 hCoV-19/England/ALDP-BBA143/2020|EPI_ISL_701997|20 ···················· ···················· ····················· ···················· ···················· ····················
W 5770 hCoV-19/England/ALDP-BBB0DC/2020|EPI_ISL_701998|20 ···················· ···················· ····················· ···················· ···················· ····················
W 5771 hCoV-19/England/ALDP-BB37A2/2020|EPI_ISL_701999|20 ···················· ···················· ····················· ···················· ···················· ····················
W 5772 hCoV-19/England/QEUH-BCB9B0/2020|EPI_ISL_704191|20 ···················· ···················· ····················· ···················· ···················· ····················
W 5773 hCoV-19/England/QEUH-BC37BE/2020|EPI_ISL_704192|20 ···················· ···················· ····················· ···················· ···················· ····················
W 5774 hCoV-19/England/QEUH-BCC120/2020|EPI_ISL_704197|20 ···················· ···················· ····················· ···················· ···················· ····················
W 5775 hCoV-19/England/QEUH-BCC6E5/2020|EPI_ISL_704199|20 ···················· ···················· ····················· ···················· ···················· ····················
W 5776 hCoV-19/Wales/PHWC-491F7C/2020|EPI_ISL_706800|2020 ···················· ···················· ····················· ···················· ···················· ····················
W 5780 hCoV-19/Wales/PHWC-491FD6/2020|EPI_ISL_706804|2020 ···················· ···················· ····················· ···················· ···················· ····················
W 5781 hCoV-19/Wales/PHWC-491FE5/2020|EPI_ISL_706805|2020 ···················· ···················· ····················· ···················· ···················· ····················
W 5782 hCoV-19/Wales/PHWC-491FF4/2020|EPI_ISL_706806|2020 ···················· ···················· ····················· ···················· ···················· ····················
W 5783 hCoV-19/Wales/PHWC-49200F/2020|EPI_ISL_706807|2020 ···················· ···················· ····················· ···················· ···················· ····················
W 5784 hCoV-19/Wales/PHWC-49201E/2020|EPI_ISL_706808|2020 ···················· ···················· ····················· ···················· ···················· ····················
W 5785 hCoV-19/Wales/PHWC-49202D/2020|EPI_ISL_706809|2020 ···················· ···················· ····················· ···················· ···················· ····················
W 5787 hCoV-19/Wales/PHWC-4921FD/2020|EPI_ISL_706830|2020 ···················· ···················· ····················· ···················· ···················· ····················
W 5789 hCoV-19/Wales/PHWC-492263/2020|EPI_ISL_706832|2020 ···················· ···················· ····················· ···················· ···················· ····················
W 5790 hCoV-19/Wales/PHWC-492272/2020|EPI_ISL_706833|2020 ···················· ···················· ····················· ···················· ···················· ····················
W 5791 hCoV-19/Wales/PHWC-492281/2020|EPI_ISL_706834|2020 ···················· ···················· ····················· ···················· ···················· ····················
W 5792 hCoV-19/Wales/PHWC-492290/2020|EPI_ISL_706835|2020 ···················· ···················· ····················· ···················· ···················· ····················
W 5794 hCoV-19/Wales/PHWC-4922DC/2020|EPI_ISL_706837|2020 ···················· ···················· ····················· ···················· ···················· ····················
W 5795 hCoV-19/Wales/PHWC-4922FA/2020|EPI_ISL_706838|2020 ···················· ···················· ····················· ···················· ···················· ····················
W 5796 hCoV-19/Wales/PHWC-492315/2020|EPI_ISL_706839|2020 ···················· ···················· ····················· ···················· ···················· ····················
W 5800 hCoV-19/Wales/PHWC-4923E8/2020|EPI_ISL_706843|2020 ···················· ···················· ····················· ···················· ···················· ····················
W 5802 hCoV-19/Wales/PHWC-492403/2020|EPI_ISL_706845|2020 ···················· ···················· ····················· ···················· ···················· ····················
W 5803 hCoV-19/Wales/PHWC-492421/2020|EPI_ISL_706846|2020 ···················· ···················· ····················· ···················· ···················· ····················
W 5804 hCoV-19/Wales/PHWC-492430/2020|EPI_ISL_706847|2020 ···················· ···················· ····················· ···················· ···················· ····················
W 5805 hCoV-19/Wales/PHWC-49244F/2020|EPI_ISL_706848|2020 ···················· ···················· ····················· ···················· ···················· ····················
W 5806 hCoV-19/Wales/PHWC-49245E/2020|EPI_ISL_706849|2020 ···················· ···················· ····················· ···················· ···················· ····················
W 5807 hCoV-19/England/CAMC-BFB692/2020|EPI_ISL_709000|20 ···················· ···················· ····················· ···················· ···················· ····················
W 5808 hCoV-19/England/CAMC-BFB638/2020|EPI_ISL_709001|20 ···················· ···················· ····················· ···················· ···················· ····················
W 5809 hCoV-19/England/QEUH-BF0B34/2020|EPI_ISL_709005|20 ···················· ···················· ····················· ···················· ···················· ····················
W 5810 hCoV-19/England/CAMC-BFB87E/2020|EPI_ISL_709008|20 ···················· ···················· ····················· ···················· ···················· ····················
W 5811 hCoV-19/England/QEUH-BF0A0A/2020|EPI_ISL_709009|20 ···················· ···················· ····················· ···················· ···················· ····················
W 5812 hCoV-19/England/QEUH-BF0C40/2020|EPI_ISL_709013|20 ···················· ···················· ····················· ···················· ···················· ····················
W 5813 hCoV-19/England/CAMC-BFB42F/2020|EPI_ISL_709018|20 ···················· ·A·················· ····················· ···················· ···················· ···A················
W 5814 hCoV-19/England/CAMC-BFB2AD/2020|EPI_ISL_709019|20 ···················· ···················· ····················· ···················· ···················· ····················
W 5815 hCoV-19/England/CAMC-BFB1A0/2020|EPI_ISL_709021|20 ···················· ···················· ····················· ···················· ···················· ····················
W 5816 hCoV-19/England/QEUH-BF09B2/2020|EPI_ISL_709022|20 ···················· ···················· ····················· ···················· ···················· ····················
W 5817 hCoV-19/England/CAMC-BFB322/2020|EPI_ISL_709026|20 ···················· ···················· ····················· ···················· ···················· ····················
W 5818 hCoV-19/England/CAMC-BFBFA6/2020|EPI_ISL_709027|20 ···················· ···················· ····················· ···················· ···················· ····················
W 5819 hCoV-19/England/CAMC-BFB498/2020|EPI_ISL_709029|20 ···················· ···················· ····················· ···················· ···················· ····················
W 5820 hCoV-19/England/CAMC-BFB708/2020|EPI_ISL_709030|20 ···················· ···················· ····················· ···················· ···················· ····················
W 5821 hCoV-19/England/QEUH-BF0CD7/2020|EPI_ISL_709036|20 ···················· ···················· ····················· ···················· ···················· ····················
W 5822 hCoV-19/England/CAMC-BFB0EF/2020|EPI_ISL_709037|20 ···················· ···················· ····················· ···················· ···················· ····················
W 5823 hCoV-19/England/CAMC-BFB28F/2020|EPI_ISL_709038|20 ···················· ···················· ····················· ···················· ···················· ····················
W 5824 hCoV-19/England/CAMC-BF475A/2020|EPI_ISL_709039|20 ···················· ···················· ····················· ···················· ···················· ····················
W 5825 hCoV-19/England/QEUH-BF0C5F/2020|EPI_ISL_709040|20 ···················· ···················· ····················· ···················· ···················· ····················
W 5826 hCoV-19/England/QEUH-BF0A37/2020|EPI_ISL_709050|20 ·····G·············· ···················· ····················· ···················· ···················· ····················
W 5827 hCoV-19/England/CAMC-BFBB66/2020|EPI_ISL_709053|20 ···················· ···················· ····················· ···················· ···················· ····················
W 5828 hCoV-19/England/QEUH-BF09D0/2020|EPI_ISL_709056|20 ···················· ·A·················· ····················· ···················· ···················· ···A················
W 5829 hCoV-19/England/CAMC-BFBF2E/2020|EPI_ISL_709057|20 ···················· ···················· ····················· ···················· ···················· ····················
W 5830 hCoV-19/England/QEUH-BF0BE9/2020|EPI_ISL_709058|20 ···················· ···················· ····················· ···················· ···················· ····················
W 5831 hCoV-19/England/QEUH-BF0ADD/2020|EPI_ISL_709060|20 ···················· ···················· ····················· ···················· ···················· ····················
W 5832 hCoV-19/England/CAMC-BFB489/2020|EPI_ISL_709061|20 ···················· ···················· ····················· ···················· ···················· ····················
W 5833 hCoV-19/England/CAMC-BFB50E/2020|EPI_ISL_709062|20 ···················· ·A·················· ····················· ···················· ···················· ····················
W 5834 hCoV-19/England/CAMC-BFBF97/2020|EPI_ISL_709063|20 ···················· ···················· ····················· ···················· ······T············· ····················
W 5835 hCoV-19/England/CAMC-BFBEC7/2020|EPI_ISL_709064|20 ···················· ·A·················· ····················· ···················· ···················· ····················
W 5836 hCoV-19/England/CAMC-BFBD42/2020|EPI_ISL_709070|20 ···················· ···················· ····················· ···················· ···················· ····················
W 5837 hCoV-19/England/CAMC-BFB753/2020|EPI_ISL_709072|20 ···················· ···················· ····················· ···················· ···················· ····················
W 5838 hCoV-19/England/CAMC-BFB51D/2020|EPI_ISL_709075|20 ···················· ·A·················· ····················· ···················· ···················· ····················
W 5839 hCoV-19/England/QEUH-BF0AFB/2020|EPI_ISL_709077|20 ···················· ···················· ····················· ···················· ···················· ····················
W 5840 hCoV-19/England/QEUH-BF0CF5/2020|EPI_ISL_709078|20 ···················· ···················· ····················· ···················· ···················· ····················

| | | | | | | | |
|---|---|---|---|---|---|---|---|
| W | 5841 hCoV-19/England/QEUH-BF0A28/2020\|EPI_ISL_709079\|20 | ···················· | ···················· | ···················· | ···················· | ···················· | ···················· |
| W | 5842 hCoV-19/England/CAMC-BFB8AB/2020\|EPI_ISL_709081\|20 | ···················· | ···················· | ···················· | ···················· | ···················· | ···················· |
| W | 5843 hCoV-19/England/QEUH-BF0A73/2020\|EPI_ISL_709082\|20 | ···················· | ···················· | ···················· | ···················· | ···················· | ···················· |
| W | 5844 hCoV-19/England/CAMC-BFB717/2020\|EPI_ISL_709088\|20 | ···················· | ···················· | ···················· | ···················· | ···················· | ···················· |
| W | 5845 hCoV-19/Gibraltar/205000674/2020\|EPI_ISL_710000\|20 | ···················· | ···················· | ···················· | ···················· | ···················· | ···················· |
| W | 5846 hCoV-19/Gibraltar/205000675/2020\|EPI_ISL_710001\|20 | ···················· | ···················· | ···················· | ···················· | ···················· | ···················· |
| W | 5847 hCoV-19/Gibraltar/205000676/2020\|EPI_ISL_710002\|20 | ···················· | ···················· | ···················· | ···················· | ···················· | ···················· |
| W | 5848 hCoV-19/Gibraltar/205000677/2020\|EPI_ISL_710003\|20 | ···················· | ···················· | ···················· | ···················· | ···················· | ···················· |
| W | 5849 hCoV-19/Gibraltar/205000679/2020\|EPI_ISL_710004\|20 | ···················· | ···················· | ···················· | ···················· | ···················· | ···················· |
| W | 5850 hCoV-19/Gibraltar/205000680/2020\|EPI_ISL_710005\|20 | ···················· | ···················· | ···················· | ···················· | ···················· | ···················· |
| W | 5851 hCoV-19/Gibraltar/205000681/2020\|EPI_ISL_710006\|20 | ···················· | ···················· | ···················· | ···················· | ···················· | ···············A···· |
| W | 5852 hCoV-19/Gibraltar/205000683/2020\|EPI_ISL_710007\|20 | ···················· | ···················· | ···················· | ···················· | ···················· | ···················· |
| W | 5853 hCoV-19/Gibraltar/205000684/2020\|EPI_ISL_710008\|20 | ···················· | ···················· | ···················· | ···················· | ···················· | ···················· |
| W | 5854 hCoV-19/Gibraltar/205000685/2020\|EPI_ISL_710009\|20 | ···················· | ···················· | ···················· | ···················· | ···················· | ···················· |
| W | 5855 hCoV-19/Gibraltar/205000686/2020\|EPI_ISL_710010\|20 | ···················· | ···················· | ···················· | ···················· | ···················· | ···················· |
| W | 5856 hCoV-19/Gibraltar/205000687/2020\|EPI_ISL_710011\|20 | ···················· | ···················· | ···················· | ···················· | ···················· | ···················· |
| W | 5857 hCoV-19/Gibraltar/205000688/2020\|EPI_ISL_710012\|20 | ···················· | ···················· | ···················· | ···················· | ···················· | ···················· |
| W | 5858 hCoV-19/Gibraltar/205000689/2020\|EPI_ISL_710013\|20 | ···················· | ···················· | ···················· | ···················· | ···················· | ···················· |
| W | 5859 hCoV-19/Gibraltar/205000690/2020\|EPI_ISL_710014\|20 | ···················· | ···················· | ···················· | ···················· | ···················· | ···················· |
| W | 5860 hCoV-19/Gibraltar/205000691/2020\|EPI_ISL_710015\|20 | ···················· | ···················· | ···················· | ···················· | ···················· | ···················· |
| W | 5861 hCoV-19/Gibraltar/205000692/2020\|EPI_ISL_710016\|20 | ···················· | ···················· | ···················· | ···················· | ···················· | ···············A···· |
| W | 5862 hCoV-19/Gibraltar/205000693/2020\|EPI_ISL_710017\|20 | ···················· | ···················· | ···················· | ···················· | ···················· | ···················· |
| W | 5863 hCoV-19/Gibraltar/205000694/2020\|EPI_ISL_710018\|20 | ···················· | ···················· | ···················· | ···················· | ···················· | ···················· |
| W | 5864 hCoV-19/Gibraltar/205000695/2020\|EPI_ISL_710019\|20 | ···················· | ···················· | ···················· | ···················· | ···················· | ···················· |
| W | 5865 hCoV-19/Gibraltar/205000696/2020\|EPI_ISL_710020\|20 | ···················· | ···················· | ···················· | ···················· | ···················· | ···················· |
| W | 5866 hCoV-19/Gibraltar/205000697/2020\|EPI_ISL_710021\|20 | ···················· | ···················· | ···················· | ···················· | ···················· | ···················· |
| W | 5867 hCoV-19/Gibraltar/205000698/2020\|EPI_ISL_710022\|20 | ···················· | ·A·················· | ···················· | ···················· | ···················· | ···················· |
| W | 5868 hCoV-19/Gibraltar/205000699/2020\|EPI_ISL_710023\|20 | ···················· | ···················· | ···················· | ···················· | ···················· | ···················· |
| W | 5869 hCoV-19/Gibraltar/205000701/2020\|EPI_ISL_710024\|20 | ···················· | ···················· | ···················· | ···················· | ···················· | ···················· |
| W | 5870 hCoV-19/Gibraltar/205000703/2020\|EPI_ISL_710025\|20 | ···················· | ···················· | ···················· | ···················· | ···················· | ···················· |
| W | 5871 hCoV-19/Gibraltar/205000704/2020\|EPI_ISL_710026\|20 | ···················· | ···················· | ···················· | ···················· | ···················· | ···················· |
| W | 5872 hCoV-19/Gibraltar/205000705/2020\|EPI_ISL_710027\|20 | ···················· | ·A·················· | ···················· | ···················· | ···················· | ···················· |
| W | 5873 hCoV-19/Gibraltar/205000706/2020\|EPI_ISL_710028\|20 | ···················· | ···················· | ···················· | ···················· | ···················· | ···················· |
| W | 5874 hCoV-19/Gibraltar/205000707/2020\|EPI_ISL_710029\|20 | ···················· | ···················· | ···················· | ···················· | ···················· | ···················· |
| W | 5875 hCoV-19/Gibraltar/205000708/2020\|EPI_ISL_710030\|20 | ···················· | ···················· | ···················· | ···················· | ···················· | ···················· |
| W | 5876 hCoV-19/Gibraltar/205000709/2020\|EPI_ISL_710031\|20 | ···················· | ···················· | ···················· | ···················· | ···················· | ···················· |
| W | 5877 hCoV-19/Gibraltar/205000710/2020\|EPI_ISL_710032\|20 | ···················· | ·A·················· | ···················· | ···················· | ···················· | ··A················· |
| W | 5878 hCoV-19/Gibraltar/205000711/2020\|EPI_ISL_710033\|20 | ···················· | ···················· | ···················· | ···················· | ···················· | ···················· |
| W | 5879 hCoV-19/USA/MD-MDH-0477/2020\|EPI_ISL_710090\|2020-1 | ···················· | ···················· | ···················· | ···················· | ···················· | ···················· |
| W | 5880 hCoV-19/USA/MD-MDH-0471/2020\|EPI_ISL_710091\|2020-1 | ···················· | ···················· | ···················· | ···················· | ···················· | ···················· |
| W | 5881 hCoV-19/USA/MD-MDH-0474/2020\|EPI_ISL_710092\|2020-1 | ···················· | ···················· | ···················· | ···················· | ···················· | ···················· |
| W | 5882 hCoV-19/USA/MD-MDH-0476/2020\|EPI_ISL_710093\|2020-1 | ···················· | ···················· | ···················· | ···················· | ···················· | ···················· |
| W | 5883 hCoV-19/USA/MD-MDH-0479/2020\|EPI_ISL_710094\|2020-1 | ···················· | ···················· | ···················· | ···················· | ···················· | ···················· |
| W | 5884 hCoV-19/USA/MD-MDH-0481/2020\|EPI_ISL_710095\|2020-1 | ···················· | ···················· | ···················· | ···················· | ···················· | ···················· |
| W | 5885 hCoV-19/USA/MD-MDH-0482/2020\|EPI_ISL_710096\|2020-1 | ···················· | ···················· | ···················· | ···················· | ···················· | ···················· |
| W | 5886 hCoV-19/USA/MD-MDH-0483/2020\|EPI_ISL_710097\|2020-1 | ···················· | ·A·················· | ···················· | ···················· | ···················· | ···················· |
| W | 5887 hCoV-19/USA/MD-MDH-0484/2020\|EPI_ISL_710098\|2020-1 | ···················· | ·A·················· | ···················· | ···················· | ···················· | ···················· |
| W | 5888 hCoV-19/USA/CA-LACPHL-AA00001/2020\|EPI_ISL_710099\| | ···················· | ···················· | ···················· | ···················· | ···················· | ···················· |
| W | 5889 hCoV-19/United Arab Emirates/0324/2020\|EPI_ISL_698 | ···················· | ···················· | ···················· | ···················· | ···················· | ···················· |
| W | 5890 hCoV-19/United Arab Emirates/0325/2020\|EPI_ISL_698 | ···················· | ··········T········· | ···················· | ···················· | ···················· | ···················· |
| W | 5891 hCoV-19/United Arab Emirates/0326/2020\|EPI_ISL_698 | ···················· | ···················· | ···················· | ···················· | ···················· | ···················· |
| W | 5892 hCoV-19/United Arab Emirates/0327/2020\|EPI_ISL_698 | ···················· | ···················· | ···················· | ···················· | ···················· | ···················· |
| W | 5893 hCoV-19/United Arab Emirates/0328/2020\|EPI_ISL_698 | ···················· | ···················· | ···················· | ···················· | ···················· | ···················· |
| W | 5894 hCoV-19/United Arab Emirates/0329/2020\|EPI_ISL_698 | ···················· | ···················· | ···················· | ···················· | ···················· | ···················· |
| W | 5895 hCoV-19/United Arab Emirates/0330/2020\|EPI_ISL_698 | ···················· | ···················· | ···················· | ···················· | ···················· | ···················· |
| W | 5896 hCoV-19/United Arab Emirates/0331/2020\|EPI_ISL_698 | ···················· | ···················· | ···················· | ···················· | ···················· | ···················· |
| W | 5897 hCoV-19/United Arab Emirates/0332/2020\|EPI_ISL_698 | ···················· | ···················· | ···················· | ···················· | ···················· | ···················· |
| W | 5898 hCoV-19/United Arab Emirates/0333/2020\|EPI_ISL_698 | ···················· | ···················· | ···················· | ···················· | ···················· | ···················· |
| W | 5899 hCoV-19/United Arab Emirates/0334/2020\|EPI_ISL_698 | ···················· | ···················· | ···················· | ···················· | ···················· | ···················· |
| W | 5900 hCoV-19/United Arab Emirates/0335/2020\|EPI_ISL_698 | ···················· | ···················· | ···················· | ···················· | ···················· | ···················· |
| W | 5901 hCoV-19/United Arab Emirates/0336/2020\|EPI_ISL_698 | ···················· | ···················· | ···················· | ···················· | ···················· | ···················· |
| W | 5902 hCoV-19/United Arab Emirates/0338/2020\|EPI_ISL_698 | ···················· | ···················· | ···················· | ···················· | ···················· | ···················· |
| W | 5903 hCoV-19/United Arab Emirates/0342/2020\|EPI_ISL_698 | ···················· | ···················· | ···················· | ···················· | ···················· | ···················· |
| W | 5904 hCoV-19/United Arab Emirates/0343/2020\|EPI_ISL_698 | ···················· | ···················· | ···················· | ···················· | ···················· | ···················· |
| W | 5905 hCoV-19/United Arab Emirates/0344/2020\|EPI_ISL_698 | ···················· | ···················· | ···················· | ···················· | ···················· | ···················· |
| W | 5906 hCoV-19/United Arab Emirates/0404/2020\|EPI_ISL_698 | ···················· | ···················· | ···················· | ···················· | ···················· | ···················· |
| W | 5907 hCoV-19/United Arab Emirates/0405/2020\|EPI_ISL_698 | ···················· | ···················· | ···················· | ···················· | ···················· | ···················· |
| W | 5908 hCoV-19/United Arab Emirates/0406/2020\|EPI_ISL_698 | ···················· | ···················· | ···················· | ···················· | ···················· | ···················· |
| W | 5909 hCoV-19/United Arab Emirates/0407/2020\|EPI_ISL_698 | ···················· | ···················· | ···················· | ···················· | ···················· | ···················· |
| W | 5910 hCoV-19/United Arab Emirates/0408/2020\|EPI_ISL_698 | ···················· | ···················· | ···················· | ···················· | ···················· | ···················· |
| W | 5911 hCoV-19/United Arab Emirates/0409/2020\|EPI_ISL_698 | ···················· | ···················· | ···················· | ···················· | ···················· | ···················· |
| W | 5912 hCoV-19/United Arab Emirates/0410/2020\|EPI_ISL_698 | ···················· | ···················· | ···················· | ···················· | ···················· | ···················· |
| W | 5913 hCoV-19/United Arab Emirates/0411/2020\|EPI_ISL_698 | ···················· | ···················· | ···················· | ···················· | ···················· | ···················· |
| W | 5914 hCoV-19/United Arab Emirates/0412/2020\|EPI_ISL_698 | ···················· | ···················· | ···················· | ···················· | ···················· | ···················· |
| W | 5915 hCoV-19/United Arab Emirates/0413/2020\|EPI_ISL_698 | ···················· | ···················· | ···················· | ···················· | ···················· | ···················· |
| W | 5916 hCoV-19/United Arab Emirates/0414/2020\|EPI_ISL_698 | ···················· | ···················· | ···················· | ···················· | ···················· | ···················· |
| W | 5917 hCoV-19/United Arab Emirates/0415/2020\|EPI_ISL_698 | ···················· | ···················· | ···················· | ···················· | ···················· | ···················· |
| W | 5918 hCoV-19/United Arab Emirates/0416/2020\|EPI_ISL_698 | ···················· | ···················· | ···················· | ···················· | ···················· | ···················· |
| W | 5919 hCoV-19/United Arab Emirates/0417/2020\|EPI_ISL_698 | ···················· | ···················· | ···················· | ···················· | ···················· | ···················· |
| W | 5920 hCoV-19/United Arab Emirates/0418/2020\|EPI_ISL_698 | ···················· | ···················· | ···················· | ···················· | ···················· | ···················· |

| | | | | | | | | | |
|---|---|---|---|---|---|---|---|---|---|
| W | 5921 | hCoV-19/United Arab Emirates/0419/2020\|EPI_ISL_698 | ···················· | ···················· | ···················· | ···················· | ···················· | ···················· | |
| W | 5922 | hCoV-19/United Arab Emirates/0420/2020\|EPI_ISL_698 | ···················· | ···················· | ···················· | ···················· | ···················· | ···················· | |
| W | 5923 | hCoV-19/United Arab Emirates/0422/2020\|EPI_ISL_698 | ···················· | ···················· | ···················· | ···················· | ···················· | ···················· | |
| W | 5924 | hCoV-19/United Arab Emirates/0423/2020\|EPI_ISL_698 | ···················· | ···················· | ···················· | ···················· | ···················· | ···················· | |
| W | 5925 | hCoV-19/United Arab Emirates/0424/2020\|EPI_ISL_698 | ···················· | ···················· | ···················· | ···················· | ···················· | ···················· | |
| W | 5926 | hCoV-19/United Arab Emirates/0425/2020\|EPI_ISL_698 | ···················· | ···················· | ···················· | ···················· | ···················· | ···················· | |
| W | 5927 | hCoV-19/India/MH-ACTREC-144/2020\|EPI_ISL_699800\|20 | ···················· | ···················· | ···················· | ···················· | ···················· | ···················· | |
| W | 5928 | hCoV-19/India/MH-ACTREC-145/2020\|EPI_ISL_699801\|20 | ···················· | ···················· | ···················· | ···················· | ···················· | ···················· | |
| W | 5929 | hCoV-19/India/MH-ACTREC-146/2020\|EPI_ISL_699802\|20 | ···················· | ···················· | ···················· | ···················· | ···················· | ···················· | |
| W | 5930 | hCoV-19/India/MH-ACTREC-147/2020\|EPI_ISL_699803\|20 | ···················· | ···················· | ···················· | ···················· | ···················· | ···················· | |
| W | 5932 | hCoV-19/India/MH-ACTREC-149/2020\|EPI_ISL_699805\|20 | ···················· | ···················· | ···················· | ···················· | ···················· | ···················· | |
| W | 5933 | hCoV-19/India/MH-ACTREC-150/2020\|EPI_ISL_699806\|20 | ···················· | ···················· | ···················· | ···················· | ···················· | ···················· | |
| W | 5935 | hCoV-19/India/MH-ACTREC-152/2020\|EPI_ISL_699808\|20 | ···················· | ···················· | ···················· | ···················· | ···················· | ···················· | |
| W | 5936 | hCoV-19/India/MH-ACTREC-153/2020\|EPI_ISL_699809\|20 | ···················· | ···················· | ···················· | ···················· | ···················· | ···················· | |
| W | 5938 | hCoV-19/India/MH-ACTREC-156/2020\|EPI_ISL_699812\|20 | ···················· | ···················· | ···················· | ···················· | ···················· | ···················· | |
| W | 5942 | hCoV-19/India/MH-ACTREC-162/2020\|EPI_ISL_699818\|20 | ···················· | ···················· | ···················· | ···················· | ···················· | ···················· | |
| W | 5943 | hCoV-19/India/MH-ACTREC-163/2020\|EPI_ISL_699819\|20 | ···················· | ···················· | ···················· | ···················· | ···················· | ···················· | |
| W | 5944 | hCoV-19/India/MH-ACTREC-166/2020\|EPI_ISL_699822\|20 | ···················· | ···················· | ···················· | ···················· | ···················· | ···················· | |
| W | 5945 | hCoV-19/India/MH-ACTREC-167/2020\|EPI_ISL_699823\|20 | ···················· | ···················· | ···················· | ···················· | ···················· | ···················· | |
| W | 5947 | hCoV-19/India/MH-ACTREC-171/2020\|EPI_ISL_699826\|20 | ···················· | ···················· | ···················· | ···················· | ···················· | ···················· | |
| W | 5950 | hCoV-19/India/MH-ACTREC-175/2020\|EPI_ISL_699830\|20 | ···················· | ···················· | ···················· | ···················· | ···················· | ···················· | |
| W | 5952 | hCoV-19/India/MH-ACTREC-178/2020\|EPI_ISL_699833\|20 | ···················· | ················T··· | ···················· | ···················· | ···················· | ···················· | |
| W | 5954 | hCoV-19/India/MH-ACTREC-180/2020\|EPI_ISL_699835\|20 | ···················· | ···················· | ···················· | ···················· | ···················· | ···················· | |
| W | 5955 | hCoV-19/India/MH-ACTREC-183/2020\|EPI_ISL_699838\|20 | ···················· | ···················· | ···················· | ···················· | ···················· | ···················· | |
| W | 5956 | hCoV-19/India/MH-ACTREC-184/2020\|EPI_ISL_699839\|20 | ···················· | ···················· | ···················· | ···················· | ···················· | ···················· | |
| W | 5957 | hCoV-19/India/MH-ACTREC-185/2020\|EPI_ISL_699840\|20 | ···················· | ···················· | ···················· | ···················· | ···················· | ···················· | |
| W | 5958 | hCoV-19/India/MH-ACTREC-186/2020\|EPI_ISL_699841\|20 | ···················· | ···················· | ···················· | ···················· | ···················· | ···················· | |
| W | 5960 | hCoV-19/India/MH-ACTREC-192/2020\|EPI_ISL_699846\|20 | ···················· | ···················· | ···················· | ···················· | ···················· | ···················· | |
| W | 5961 | hCoV-19/India/MH-ACTREC-194/2020\|EPI_ISL_699848\|20 | ···················· | ·A·················· | ···················· | ···················· | ···················· | ···A················ | |
| W | 5962 | hCoV-19/India/MH-ACTREC-195/2020\|EPI_ISL_699849\|20 | ···················· | ·A·················· | ···················· | ···················· | ···················· | NN·A················ | 1 |
| W | 5963 | hCoV-19/England/QEUH-BCBA08/2020\|EPI_ISL_704200\|20 | ···················· | ···················· | ···················· | ···················· | ···················· | ···················· | |
| W | 5964 | hCoV-19/England/QEUH-BCBFEB/2020\|EPI_ISL_704202\|20 | ···················· | ···················· | ···················· | ···················· | ···················· | ···················· | |
| W | 5965 | hCoV-19/England/QEUH-BC3806/2020\|EPI_ISL_704204\|20 | ···················· | ···················· | ···················· | ···················· | ···················· | ···A················ | |
| W | 5966 | hCoV-19/England/QEUH-BC3851/2020\|EPI_ISL_704205\|20 | ···················· | ···················· | ···················· | ···················· | ···················· | ···················· | |
| W | 5967 | hCoV-19/England/QEUH-BC3D70/2020\|EPI_ISL_704208\|20 | ···················· | ···················· | ···················· | ···················· | ···················· | ···················· | |
| W | 5968 | hCoV-19/England/QEUH-BC3B0D/2020\|EPI_ISL_704210\|20 | ···················· | ···················· | ···················· | ···················· | ···················· | ···················· | |
| W | 5969 | hCoV-19/England/QEUH-BCCD95/2020\|EPI_ISL_704211\|20 | ···················· | ···················· | ···················· | ···················· | ···················· | ···················· | |
| W | 5970 | hCoV-19/England/QEUH-BCE513/2020\|EPI_ISL_704212\|20 | ···················· | ···················· | ···················· | ···················· | ···················· | ···················· | |
| W | 5971 | hCoV-19/England/QEUH-BCD990/2020\|EPI_ISL_704213\|20 | ···················· | ·A·················· | ···················· | ···················· | ···················· | ···A················ | |
| W | 5972 | hCoV-19/England/QEUH-BCDE0A/2020\|EPI_ISL_704214\|20 | ···················· | ···················· | ···················· | ···················· | ···················· | ···················· | |
| W | 5973 | hCoV-19/England/QEUH-BCD611/2020\|EPI_ISL_704215\|20 | ···················· | ···················· | ···················· | ···················· | ······T············· | ···················· | |
| W | 5974 | hCoV-19/England/QEUH-BCCEFC/2020\|EPI_ISL_704216\|20 | ···················· | ···················· | ···················· | ···················· | ······T············· | ···················· | |
| W | 5975 | hCoV-19/England/QEUH-BCB4FB/2020\|EPI_ISL_704220\|20 | ···················· | ···················· | ···················· | ···················· | ···················· | ···················· | |
| W | 5976 | hCoV-19/England/QEUH-BCB7B6/2020\|EPI_ISL_704221\|20 | ···················· | ···················· | ···················· | ···················· | ···················· | ···················· | |
| W | 5977 | hCoV-19/England/QEUH-BCBE1B/2020\|EPI_ISL_704223\|20 | ···················· | ···················· | ···················· | ···················· | ···················· | ···················· | |
| W | 5978 | hCoV-19/England/QEUH-BCC24B/2020\|EPI_ISL_704224\|20 | ···················· | ···················· | ···················· | ···················· | ···················· | ···················· | |
| W | 5979 | hCoV-19/England/QEUH-BCDD3A/2020\|EPI_ISL_704226\|20 | ···················· | ···················· | ···················· | ···················· | ···················· | ···················· | |
| W | 5980 | hCoV-19/England/QEUH-BCCDE0/2020\|EPI_ISL_704227\|20 | ···················· | ···················· | ···················· | ···················· | ···················· | ···················· | |
| W | 5981 | hCoV-19/England/QEUH-BCCCF2/2020\|EPI_ISL_704229\|20 | ···················· | ···················· | ···················· | ···················· | ···················· | ···················· | |
| W | 5982 | hCoV-19/England/QEUH-BCD80C/2020\|EPI_ISL_704232\|20 | ···················· | ···················· | ···················· | ···················· | ···················· | ···················· | |
| W | 5983 | hCoV-19/England/QEUH-BCCA8F/2020\|EPI_ISL_704234\|20 | ···················· | ···················· | ···················· | ···················· | ···················· | ···················· | |
| W | 5984 | hCoV-19/England/QEUH-BCDDB2/2020\|EPI_ISL_704235\|20 | ···················· | ···················· | ···················· | ···················· | ···················· | ···················· | |
| W | 5985 | hCoV-19/England/QEUH-BCD453/2020\|EPI_ISL_704240\|20 | ···················· | ···················· | ···················· | ···················· | ···················· | ···A················ | |
| W | 5986 | hCoV-19/England/QEUH-BCBD4B/2020\|EPI_ISL_704242\|20 | ···················· | ···················· | ···················· | ···················· | ···················· | ···················· | |
| W | 5987 | hCoV-19/England/QEUH-BCD198/2020\|EPI_ISL_704244\|20 | ···················· | ···················· | ···················· | ···················· | ······T············· | ···················· | |
| W | 5988 | hCoV-19/England/QEUH-BCD4EA/2020\|EPI_ISL_704245\|20 | ···················· | ···················· | ···················· | ···················· | ······T············· | ···················· | |
| W | 5989 | hCoV-19/England/QEUH-BCB4DD/2020\|EPI_ISL_704248\|20 | ···················· | ···················· | ···················· | ···················· | ···················· | ···················· | |
| W | 5990 | hCoV-19/England/QEUH-BCE258/2020\|EPI_ISL_704249\|20 | ···················· | ···················· | ···················· | ···················· | ···················· | ···················· | |
| W | 5991 | hCoV-19/England/QEUH-BCE531/2020\|EPI_ISL_704251\|20 | ···················· | ···················· | ···················· | ···················· | ···················· | ···················· | |
| W | 5992 | hCoV-19/England/QEUH-BCD73C/2020\|EPI_ISL_704253\|20 | ···················· | ···················· | ···················· | ···················· | ···················· | ···················· | |
| W | 5993 | hCoV-19/England/QEUH-BCD7E1/2020\|EPI_ISL_704254\|20 | ···················· | ···················· | ···················· | ···················· | ···················· | ···················· | |
| W | 5994 | hCoV-19/England/QEUH-BCBBC9/2020\|EPI_ISL_704261\|20 | ···················· | ···················· | ···················· | ···················· | ···················· | ···················· | |
| W | 5995 | hCoV-19/England/QEUH-BCC1C6/2020\|EPI_ISL_704263\|20 | ···················· | ···················· | ···················· | ···················· | ···················· | ···················· | |
| W | 5996 | hCoV-19/England/QEUH-BCB4EC/2020\|EPI_ISL_704264\|20 | ···················· | ···················· | ···················· | ···················· | ······T············· | ···················· | |
| W | 5997 | hCoV-19/England/QEUH-BCBAAE/2020\|EPI_ISL_704268\|20 | ···················· | ···················· | ···················· | ···················· | ···················· | ···················· | |
| W | 5998 | hCoV-19/England/QEUH-BCCE74/2020\|EPI_ISL_704290\|20 | ···················· | ···················· | ···················· | ···················· | ···················· | ···················· | |
| W | 5999 | hCoV-19/Wales/PHWC-491C0C/2020\|EPI_ISL_705527\|2020 | ···················· | ···················· | ···················· | ···················· | ···················· | ···················· | |
| W | 6000 | hCoV-19/England/ALDP-B5D8E3/2020\|EPI_ISL_705529\|20 | ···················· | ···················· | ···················· | ···················· | ···················· | ···················· | |
| W | 6001 | hCoV-19/England/ALDP-B5D8F2/2020\|EPI_ISL_705530\|20 | ···················· | ···················· | ···················· | ···················· | ···················· | ···················· | |
| W | 6002 | hCoV-19/England/ALDP-B5D959/2020\|EPI_ISL_705531\|20 | ···················· | ···················· | ···················· | ···················· | ···················· | ···················· | |
| W | 6003 | hCoV-19/England/ALDP-B5D977/2020\|EPI_ISL_705532\|20 | ···················· | ·A·················· | ···················· | ···················· | ···················· | ···A················ | |
| W | 6004 | hCoV-19/England/ALDP-B5DA38/2020\|EPI_ISL_705533\|20 | ···················· | ···················· | ···················· | ···················· | ···················· | ···················· | |
| W | 6005 | hCoV-19/England/ALDP-B5DA92/2020\|EPI_ISL_705534\|20 | ···················· | ···················· | ···················· | ···················· | ···················· | ···A················ | |
| W | 6006 | hCoV-19/England/ALDP-B5DACF/2020\|EPI_ISL_705535\|20 | ···················· | ···················· | ···················· | ···················· | ···················· | ···················· | |
| W | 6007 | hCoV-19/England/CAMB-1B78E1/2020\|EPI_ISL_705536\|20 | ···················· | ···················· | ···················· | ···················· | ······T············· | ···················· | |
| W | 6008 | hCoV-19/England/CAMB-1B7C9A/2020\|EPI_ISL_705537\|20 | ···················· | ···················· | ···················· | ···················· | ···················· | ···················· | |
| W | 6009 | hCoV-19/England/CAMB-1B7CA9/2020\|EPI_ISL_705538\|20 | ···················· | ···················· | ···················· | ···················· | ···················· | ···················· | |
| W | 6010 | hCoV-19/England/CAMB-1B7CB8/2020\|EPI_ISL_705539\|20 | ···················· | ···················· | ···················· | ···················· | ···················· | ···················· | |
| W | 6011 | hCoV-19/England/CAMB-1B7CC7/2020\|EPI_ISL_705540\|20 | ···················· | ···················· | ···················· | ···················· | ···················· | ···················· | |
| W | 6012 | hCoV-19/England/CAMB-1B7CD6/2020\|EPI_ISL_705541\|20 | ···················· | ···················· | ···················· | ···················· | ···················· | ···················· | |

| | | | | | | | | |
|---|---|---|---|---|---|---|---|---|
| W | 6013 | hCoV-19/England/CAMB-1B7CE5/2020\|EPI_ISL_705542\|20 | ···················· | ···················· | ····················· | ···················· | ···················· | ···················· |
| W | 6014 | hCoV-19/England/CAMB-1B9120/2020\|EPI_ISL_705543\|20 | ···················· | ···················· | ····················· | ···················· | ······T············· | ···················· |
| W | 6015 | hCoV-19/England/CAMB-1B91D5/2020\|EPI_ISL_705544\|20 | ···················· | ···················· | ····················· | ···················· | ···················· | ···················· |
| W | 6016 | hCoV-19/England/CAMB-1B91E4/2020\|EPI_ISL_705545\|20 | ···················· | ···················· | ····················· | ···················· | ···················· | ···················· |
| W | 6017 | hCoV-19/England/CAMB-1B91F3/2020\|EPI_ISL_705546\|20 | ···················· | ···················· | ····················· | ···················· | ···················· | ···················· |
| W | 6018 | hCoV-19/England/CAMB-1B921E/2020\|EPI_ISL_705547\|20 | ···················· | ···················· | ····················· | ···················· | ···················· | ···················· |
| W | 6019 | hCoV-19/England/CAMB-1B922D/2020\|EPI_ISL_705548\|20 | ···················· | ···················· | ····················· | ···················· | ······T············· | ···················· |
| W | 6020 | hCoV-19/England/CAMB-1B923C/2020\|EPI_ISL_705549\|20 | ···················· | ···················· | ····················· | ···················· | ···················· | ···················· |
| W | 6021 | hCoV-19/England/CAMB-1B924B/2020\|EPI_ISL_705550\|20 | ···················· | ···················· | ····················· | ···················· | ···················· | ···················· |
| W | 6022 | hCoV-19/England/CAMB-1BC934/2020\|EPI_ISL_705551\|20 | ···················· | ···················· | ····················· | ···················· | ···················· | ···················· |
| W | 6023 | hCoV-19/England/CAMB-1BC952/2020\|EPI_ISL_705552\|20 | ···················· | ···················· | ····················· | ···················· | ···················· | ···················· |
| W | 6024 | hCoV-19/England/CAMB-1BC98F/2020\|EPI_ISL_705553\|20 | ···················· | ···················· | ····················· | ···················· | ···················· | ···················· |
| W | 6025 | hCoV-19/England/CAMB-1BC9AD/2020\|EPI_ISL_705554\|20 | ···················· | ···················· | ····················· | ···················· | ···················· | ···················· |
| W | 6026 | hCoV-19/England/CAMB-1BC9E9/2020\|EPI_ISL_705555\|20 | ···················· | ···················· | ····················· | ···················· | ···················· | ···················· |
| W | 6027 | hCoV-19/England/CAMB-1BCA04/2020\|EPI_ISL_705556\|20 | ···················· | ···················· | ····················· | ···················· | ···················· | ···················· |
| W | 6028 | hCoV-19/England/CAMB-1BCA13/2020\|EPI_ISL_705557\|20 | ···················· | ···················· | ····················· | ···················· | ···················· | ···················· |
| W | 6029 | hCoV-19/England/CAMB-1BCA31/2020\|EPI_ISL_705558\|20 | ···················· | ···················· | ····················· | ···················· | ···················· | ···················· |
| W | 6030 | hCoV-19/England/CAMB-1BCA5F/2020\|EPI_ISL_705559\|20 | ···················· | ···················· | ····················· | ···················· | ···················· | ···················· |
| W | 6031 | hCoV-19/England/CAMC-BBB634/2020\|EPI_ISL_705560\|20 | ···················· | ···················· | ····················· | ···················· | ···················· | ···················· |
| W | 6032 | hCoV-19/England/CAMC-BBB75F/2020\|EPI_ISL_705561\|20 | ···················· | ···················· | ····················· | ···················· | ···················· | ···················· |
| W | 6033 | hCoV-19/England/CAMC-BBB7AA/2020\|EPI_ISL_705562\|20 | ···················· | ···················· | ····················· | ···················· | ···················· | ···················· |
| W | 6034 | hCoV-19/England/CAMC-BBB8C5/2020\|EPI_ISL_705563\|20 | ···················· | ···················· | ····················· | ···················· | ···················· | ···················· |
| W | 6035 | hCoV-19/England/CAMC-BBB8F2/2020\|EPI_ISL_705564\|20 | ···················· | ·A·················· | ····················· | ···················· | ···················· | ···················· |
| W | 6036 | hCoV-19/England/CAMC-BBB90E/2020\|EPI_ISL_705565\|20 | ···················· | ···················· | ····················· | ···················· | ···················· | ···················· |
| W | 6037 | hCoV-19/England/CAMC-BBB91D/2020\|EPI_ISL_705566\|20 | ···················· | ···················· | ····················· | ···················· | ···················· | ···················· |
| W | 6039 | hCoV-19/England/CAMC-BBBD6C/2020\|EPI_ISL_705568\|20 | ···················· | ···················· | ····················· | ···················· | ···················· | ···················· |
| W | 6040 | hCoV-19/England/CAMC-BBBD99/2020\|EPI_ISL_705569\|20 | ···················· | ···················· | ····················· | ···················· | ···················· | ···················· |
| W | 6041 | hCoV-19/England/CAMC-BBBE0F/2020\|EPI_ISL_705570\|20 | ···················· | ···················· | ····················· | ···················· | ···················· | ···················· |
| W | 6042 | hCoV-19/England/CAMC-BBBE1E/2020\|EPI_ISL_705571\|20 | ···················· | ···················· | ····················· | ···················· | ···················· | ···················· |
| W | 6043 | hCoV-19/England/CAMC-BBBEB4/2020\|EPI_ISL_705572\|20 | ···················· | ···················· | ····················· | ···················· | ···················· | ···················· |
| W | 6044 | hCoV-19/England/CAMC-BBBEC3/2020\|EPI_ISL_705573\|20 | ···················· | ···················· | ····················· | ···················· | ···················· | ···················· |
| W | 6045 | hCoV-19/England/CAMC-BBBED2/2020\|EPI_ISL_705574\|20 | ···················· | ···················· | ····················· | ···················· | ···················· | ···················· |
| W | 6046 | hCoV-19/England/CAMC-BBBF1B/2020\|EPI_ISL_705575\|20 | ···················· | ···················· | ····················· | ···················· | ···················· | ···················· |
| W | 6048 | hCoV-19/England/CAMC-BBBF48/2020\|EPI_ISL_705577\|20 | ···················· | ···················· | ····················· | ···················· | ···················· | ···················· |
| W | 6049 | hCoV-19/England/CAMC-BBBF66/2020\|EPI_ISL_705578\|20 | ···················· | ···················· | ····················· | ···················· | ···················· | ···················· |
| W | 6050 | hCoV-19/England/CAMC-BBBF93/2020\|EPI_ISL_705579\|20 | ···················· | ·A·················· | ····················· | ···················· | ···················· | ··A················· |
| W | 6051 | hCoV-19/England/CAMC-BBBFA2/2020\|EPI_ISL_705580\|20 | ···················· | ···················· | ····················· | ···················· | ···················· | ···················· |
| W | 6052 | hCoV-19/England/CAMC-BBBFB1/2020\|EPI_ISL_705581\|20 | ···················· | ···················· | ····················· | ···················· | ···················· | ···················· |
| W | 6053 | hCoV-19/England/CAMC-BBC0CC/2020\|EPI_ISL_705582\|20 | ···················· | ···················· | ····················· | ···················· | ···················· | ···················· |
| W | 6054 | hCoV-19/England/CAMC-BBC0EA/2020\|EPI_ISL_705583\|20 | ···················· | ···················· | ····················· | ···················· | ······T············· | ···················· |
| W | 6055 | hCoV-19/England/CAMC-BBC132/2020\|EPI_ISL_705584\|20 | ···················· | ···················· | ····················· | ···················· | ······T············· | ···················· |
| W | 6056 | hCoV-19/England/CAMC-BBC150/2020\|EPI_ISL_705585\|20 | ···················· | ···················· | ····················· | ···················· | ···················· | ···················· |
| W | 6057 | hCoV-19/England/CAMC-BBC1D8/2020\|EPI_ISL_705586\|20 | ···················· | ···················· | ····················· | ···················· | ···················· | ···················· |
| W | 6058 | hCoV-19/England/CAMC-BBC1E7/2020\|EPI_ISL_705587\|20 | ···················· | ···················· | ····················· | ···················· | ···················· | ···················· |
| W | 6059 | hCoV-19/England/CAMC-BBC2D5/2020\|EPI_ISL_705588\|20 | ···················· | ···················· | ····················· | ···················· | ···················· | ···················· |
| W | 6060 | hCoV-19/England/CAMC-BBC2F3/2020\|EPI_ISL_705589\|20 | ···················· | ···················· | ····················· | ···················· | ···················· | ···················· |
| W | 6061 | hCoV-19/England/CAMC-BBC30F/2020\|EPI_ISL_705590\|20 | ···················· | ···················· | ····················· | ···················· | ···················· | ···················· |
| W | 6062 | hCoV-19/England/CAMC-BBC31E/2020\|EPI_ISL_705591\|20 | ···················· | ···················· | ····················· | ···················· | ···················· | ···················· |
| W | 6063 | hCoV-19/England/CAMC-BBC32D/2020\|EPI_ISL_705592\|20 | ···················· | ···················· | ····················· | ···················· | ···················· | ···················· |
| W | 6064 | hCoV-19/England/CAMC-BBC35A/2020\|EPI_ISL_705593\|20 | ···················· | ···················· | ····················· | ···················· | ···················· | ···················· |
| W | 6065 | hCoV-19/England/CAMC-BBC509/2020\|EPI_ISL_705594\|20 | ···················· | ···················· | ····················· | ···················· | ···················· | ···················· |
| W | 6066 | hCoV-19/England/CAMC-BBC7D6/2020\|EPI_ISL_705595\|20 | ···················· | ···················· | ····················· | ···················· | ···················· | ···················· |
| W | 6067 | hCoV-19/England/CAMC-BBC7E5/2020\|EPI_ISL_705596\|20 | ···················· | ···················· | ····················· | ···················· | ···················· | ···················· |
| W | 6068 | hCoV-19/England/CAMC-BBC7F4/2020\|EPI_ISL_705597\|20 | ···················· | ···················· | ····················· | ···················· | ···················· | ···················· |
| W | 6069 | hCoV-19/England/CAMC-BBC800/2020\|EPI_ISL_705598\|20 | ···················· | ···················· | ····················· | ···················· | ···················· | ···················· |
| W | 6070 | hCoV-19/England/CAMC-BBC82E/2020\|EPI_ISL_705599\|20 | ···················· | ···················· | ····················· | ···················· | ···················· | ···················· |
| W | 6072 | hCoV-19/Wales/PHWC-49205A/2020\|EPI_ISL_706811\|2020 | ···················· | ···················· | ····················· | ···················· | ···················· | ···················· |
| W | 6073 | hCoV-19/Wales/PHWC-492069/2020\|EPI_ISL_706812\|2020 | ···················· | ···················· | ····················· | ···················· | ···················· | ···················· |
| W | 6074 | hCoV-19/Wales/PHWC-492078/2020\|EPI_ISL_706813\|2020 | ···················· | ···················· | ····················· | ···················· | ···················· | ···················· |
| W | 6077 | hCoV-19/Wales/PHWC-4920B4/2020\|EPI_ISL_706816\|2020 | ···················· | ···················· | ····················· | ···················· | ···················· | ···················· |
| W | 6078 | hCoV-19/Wales/PHWC-4920C3/2020\|EPI_ISL_706817\|2020 | ···················· | ···················· | ····················· | ···················· | ···················· | ···················· |
| W | 6079 | hCoV-19/Wales/PHWC-4920E1/2020\|EPI_ISL_706818\|2020 | ···················· | ···················· | ····················· | ···················· | ···················· | ···················· |
| W | 6080 | hCoV-19/Wales/PHWC-4920F0/2020\|EPI_ISL_706819\|2020 | ···················· | ···················· | ····················· | ···················· | ···················· | ···················· |
| W | 6082 | hCoV-19/Wales/PHWC-49211B/2020\|EPI_ISL_706821\|2020 | ···················· | ···················· | ····················· | ···················· | ···················· | ···················· |
| W | 6083 | hCoV-19/Wales/PHWC-49212A/2020\|EPI_ISL_706822\|2020 | ···················· | ···················· | ····················· | ···················· | ···················· | ···················· |
| W | 6085 | hCoV-19/Wales/PHWC-492157/2020\|EPI_ISL_706824\|2020 | ···················· | ···················· | ····················· | ···················· | ···················· | ···················· |
| W | 6086 | hCoV-19/Wales/PHWC-492184/2020\|EPI_ISL_706825\|2020 | ···················· | ···················· | ····················· | ···················· | ···················· | ···················· |
| W | 6087 | hCoV-19/Wales/PHWC-492193/2020\|EPI_ISL_706826\|2020 | ···················· | ···················· | ····················· | ···················· | ···················· | ···················· |
| W | 6088 | hCoV-19/Wales/PHWC-4921A2/2020\|EPI_ISL_706827\|2020 | ···················· | ···················· | ····················· | ···················· | ···················· | ···················· |
| W | 6089 | hCoV-19/Wales/PHWC-4921B1/2020\|EPI_ISL_706828\|2020 | ···················· | ···················· | ····················· | ···················· | ···················· | ···················· |
| W | 6091 | hCoV-19/Wales/PHWC-4924D6/2020\|EPI_ISL_706851\|2020 | ···················· | ···················· | ····················· | ···················· | ···················· | ···················· |
| W | 6092 | hCoV-19/Wales/PHWC-4924E5/2020\|EPI_ISL_706852\|2020 | ···················· | ···················· | ····················· | ···················· | ···················· | ···················· |
| W | 6094 | hCoV-19/Wales/PHWC-49252E/2020\|EPI_ISL_706854\|2020 | ···················· | ···················· | ····················· | ···················· | ···················· | ···················· |
| W | 6095 | hCoV-19/Wales/PHWC-49253D/2020\|EPI_ISL_706855\|2020 | ···················· | ···················· | ····················· | ···················· | ···················· | ···················· |
| W | 6096 | hCoV-19/Wales/PHWC-49254C/2020\|EPI_ISL_706856\|2020 | ···················· | ···················· | ····················· | ···················· | ···················· | ···················· |
| W | 6097 | hCoV-19/Wales/PHWC-49256A/2020\|EPI_ISL_706857\|2020 | ···················· | ···················· | ····················· | ···················· | ···················· | ···················· |
| W | 6098 | hCoV-19/Wales/PHWC-492579/2020\|EPI_ISL_706858\|2020 | ···················· | ···················· | ····················· | ···················· | ···················· | ···················· |
| W | 6100 | hCoV-19/Wales/PHWC-492597/2020\|EPI_ISL_706860\|2020 | ···················· | ···················· | ····················· | ···················· | ···················· | ···················· |
| W | 6101 | hCoV-19/Wales/PHWC-4925A6/2020\|EPI_ISL_706861\|2020 | ···················· | ···················· | ····················· | ···················· | ···················· | ···················· |
| W | 6102 | hCoV-19/Wales/PHWC-4925B5/2020\|EPI_ISL_706862\|2020 | ···················· | ···················· | ····················· | ···················· | ···················· | ···················· |

| | | | | | | | |
|---|---|---|---|---|---|---|---|
| W | 6104 hCoV-19/England/PORT-2D4BD5/2020\|EPI_ISL_706864\|20 | ···················· | ···················· | ····················· | ···················· | ······T············· | ···················· |
| W | 6105 hCoV-19/England/PORT-2D4CC3/2020\|EPI_ISL_706865\|20 | ···················· | ···················· | ····················· | ···················· | ···················· | ···················· |
| W | 6106 hCoV-19/England/PORT-2D4CE1/2020\|EPI_ISL_706866\|20 | ···················· | ···················· | ····················· | ···················· | ······T············· | ···················· |
| W | 6107 hCoV-19/England/PORT-2D4D2A/2020\|EPI_ISL_706867\|20 | ···················· | ···················· | ····················· | ···················· | ···················· | ···················· |
| W | 6108 hCoV-19/England/PORT-2D4D48/2020\|EPI_ISL_706868\|20 | ···················· | ···················· | ····················· | ···················· | ···················· | ···················· |
| W | 6109 hCoV-19/England/PORT-2D4D66/2020\|EPI_ISL_706869\|20 | ···················· | ···················· | ····················· | ···················· | ···················· | ···················· |
| W | 6110 hCoV-19/England/PORT-2D4D93/2020\|EPI_ISL_706870\|20 | ···················· | ···················· | ····················· | ···················· | ······T············· | ···················· |
| W | 6111 hCoV-19/England/PORT-2D4DB1/2020\|EPI_ISL_706871\|20 | ···················· | ···················· | ····················· | ···················· | ···················· | ···················· |
| W | 6112 hCoV-19/England/PORT-2E9B64/2020\|EPI_ISL_706872\|20 | ···················· | ···················· | ····················· | ···················· | ······T············· | ···················· |
| W | 6113 hCoV-19/England/PORT-2E9BA0/2020\|EPI_ISL_706873\|20 | ···················· | ···················· | ····················· | ···················· | ···················· | ···················· |
| W | 6114 hCoV-19/England/PORT-2E9BBF/2020\|EPI_ISL_706874\|20 | ···················· | ···················· | ····················· | ···················· | ···················· | ···················· |
| W | 6115 hCoV-19/England/PORT-2E9C43/2020\|EPI_ISL_706875\|20 | ···················· | ···················· | ····················· | ···················· | ···················· | ···················· |
| W | 6116 hCoV-19/England/PORT-2E9CBC/2020\|EPI_ISL_706876\|20 | ···················· | ···················· | ····················· | ···················· | ···················· | ···················· |
| W | 6117 hCoV-19/England/PORT-2E9CCB/2020\|EPI_ISL_706877\|20 | ···················· | ···················· | ····················· | ···················· | ······T············· | ···················· |
| W | 6118 hCoV-19/England/PORT-2E9D04/2020\|EPI_ISL_706878\|20 | ···················· | ···················· | ····················· | ···················· | ···················· | ···················· |
| W | 6119 hCoV-19/England/PORT-2E9D13/2020\|EPI_ISL_706879\|20 | ···················· | ···················· | ····················· | ···················· | ···················· | ···················· |
| W | 6120 hCoV-19/England/PORT-2E9D31/2020\|EPI_ISL_706880\|20 | ···················· | ···················· | ····················· | ···················· | ······T············· | ···················· |
| W | 6121 hCoV-19/England/PORT-2E9D9B/2020\|EPI_ISL_706881\|20 | ···················· | ···················· | ····················· | ···················· | ······T············· | ···················· |
| W | 6122 hCoV-19/England/PORT-2F5066/2020\|EPI_ISL_706882\|20 | ···················· | ···················· | ····················· | ···················· | ······T············· | ···················· |
| W | 6123 hCoV-19/England/PORT-2F50C0/2020\|EPI_ISL_706883\|20 | ···················· | ···················· | ····················· | ···················· | ···················· | ···················· |
| W | 6124 hCoV-19/England/PORT-2F50EE/2020\|EPI_ISL_706884\|20 | ···················· | ···················· | ····················· | ···················· | ···················· | ···················· |
| W | 6125 hCoV-19/England/PORT-2F5118/2020\|EPI_ISL_706885\|20 | ···················· | ···················· | ····················· | ···················· | ···················· | ···················· |
| W | 6126 hCoV-19/England/PORT-2F528E/2020\|EPI_ISL_706886\|20 | ···················· | ···················· | ····················· | ···················· | ······T············· | ···················· |
| W | 6127 hCoV-19/England/PORT-2F5303/2020\|EPI_ISL_706887\|20 | ···················· | ···················· | ····················· | ···················· | ···················· | ···················· |
| W | 6128 hCoV-19/England/PORT-2F534F/2020\|EPI_ISL_706888\|20 | ···················· | ···················· | ····················· | ···················· | ···················· | ···················· |
| W | 6129 hCoV-19/England/PORT-2F55A3/2020\|EPI_ISL_706889\|20 | ···················· | ···················· | ····················· | ···················· | ······T············· | ···················· |
| W | 6130 hCoV-19/England/PORT-2F55C1/2020\|EPI_ISL_706890\|20 | ···················· | ···················· | ····················· | ···················· | ···················· | ···················· |
| W | 6131 hCoV-19/Scotland/QEUH-A35D6B/2020\|EPI_ISL_706891\|2 | ···················· | ···················· | ····················· | ···················· | ···················· | ···················· |
| W | 6132 hCoV-19/England/QEUH-A35D7A/2020\|EPI_ISL_706892\|20 | ···················· | ···················· | ····················· | ···················· | ···················· | ···················· |
| W | 6133 hCoV-19/England/QEUH-A35DB6/2020\|EPI_ISL_706893\|20 | ···················· | ···················· | ····················· | ···················· | ···················· | ···················· |
| W | 6134 hCoV-19/Scotland/QEUH-A35DC5/2020\|EPI_ISL_706894\|2 | ···················· | ···················· | ····················· | ···················· | ···················· | ···················· |
| W | 6135 hCoV-19/England/QEUH-A35DF2/2020\|EPI_ISL_706895\|20 | ···················· | ···················· | ····················· | ···················· | ···················· | ···················· |
| W | 6136 hCoV-19/England/QEUH-A35E1D/2020\|EPI_ISL_706896\|20 | ···················· | ···················· | ····················· | ···················· | ···················· | ···················· |
| W | 6137 hCoV-19/Scotland/QEUH-A35E4A/2020\|EPI_ISL_706897\|2 | ···················· | ···················· | ····················· | ···················· | ···················· | ···················· |
| W | 6138 hCoV-19/Scotland/QEUH-A35EB3/2020\|EPI_ISL_706898\|2 | ···················· | ···················· | ····················· | ···················· | ···················· | ···················· |
| W | 6139 hCoV-19/England/QEUH-A35EC2/2020\|EPI_ISL_706899\|20 | ···················· | ···················· | ····················· | ···················· | ···················· | ···················· |
| W | 6140 hCoV-19/England/QEUH-BF19FD/2020\|EPI_ISL_709091\|20 | ···················· | ···················· | ····················· | ···················· | ···················· | ···················· |
| W | 6141 hCoV-19/England/CAMC-BFB2DA/2020\|EPI_ISL_709094\|20 | ···················· | ···················· | ····················· | ···················· | ···················· | ···················· |
| W | 6142 hCoV-19/England/CAMC-BFB771/2020\|EPI_ISL_709096\|20 | ···················· | ···················· | ····················· | ···················· | ···················· | ···················· |
| W | 6143 hCoV-19/USA/TX-DSHS-1655/2020\|EPI_ISL_710087\|2020- | ···················· | ·A·················· | ····················· | ···················· | ···················· | ···················· |
| W | 6144 hCoV-19/USA/TX-DSHS-1668/2020\|EPI_ISL_710088\|2020- | ···················· | ·A·················· | ····················· | ···················· | ···················· | ···················· |
| W | 6145 hCoV-19/USA/AZ-TG408159/2020\|EPI_ISL_694911\|2020-0 | ···················· | ·A·················· | ····················· | ···················· | ···················· | ···················· |
| W | 6146 hCoV-19/USA/AZ-TG408160/2020\|EPI_ISL_694912\|2020-0 | ···················· | ·A·················· | ····················· | ···················· | ···················· | ···················· |
| W | 6147 hCoV-19/USA/AZ-TG408162/2020\|EPI_ISL_694914\|2020-0 | ···················· | ···················· | ····················· | ···················· | ···················· | ···················· |
| W | 6148 hCoV-19/USA/AZ-TG408164/2020\|EPI_ISL_694916\|2020-0 | ···················· | ···················· | ····················· | ···················· | ···················· | ···················· |
| W | 6149 hCoV-19/USA/AZ-TG408166/2020\|EPI_ISL_694917\|2020-0 | ···················· | ···················· | ····················· | ···················· | ···················· | ···················· |
| W | 6150 hCoV-19/USA/AZ-TG408181/2020\|EPI_ISL_694923\|2020-0 | ···················· | ·············A······ | ····················· | ···················· | ···················· | ···················· |
| W | 6151 hCoV-19/USA/AZ-TG408182/2020\|EPI_ISL_694924\|2020-0 | ···················· | ·············A······ | ····················· | ···················· | ···················· | ···················· |
| W | 6152 hCoV-19/USA/AZ-TG408183/2020\|EPI_ISL_694925\|2020-0 | ···················· | ·A·················· | ····················· | ···················· | ···················· | ···················· |
| W | 6153 hCoV-19/USA/AZ-TG408188/2020\|EPI_ISL_694928\|2020-0 | ···················· | ···················· | ····················· | ···················· | ···················· | ···················· |
| W | 6154 hCoV-19/USA/AZ-TG408190/2020\|EPI_ISL_694930\|2020-0 | ···················· | ···················· | ····················· | ···················· | ···················· | ···················· |
| W | 6155 hCoV-19/USA/AZ-TG408195/2020\|EPI_ISL_694933\|2020-0 | ···················· | ···················· | ····················· | ···················· | ···················· | ···················· |
| W | 6156 hCoV-19/USA/AZ-TG408196/2020\|EPI_ISL_694934\|2020-0 | ···················· | ···················· | ····················· | ···················· | ···················· | ···················· |
| W | 6157 hCoV-19/USA/AZ-TG408199/2020\|EPI_ISL_694937\|2020-0 | ···················· | ·A·················· | ····················· | ···················· | ···················· | ···················· |
| W | 6158 hCoV-19/USA/AZ-TG408203/2020\|EPI_ISL_694940\|2020-0 | ···················· | ·A·················· | ····················· | ···················· | ···················· | ···················· |
| W | 6159 hCoV-19/USA/AZ-TG408205/2020\|EPI_ISL_694941\|2020-0 | ···················· | ·············A······ | ····················· | ···················· | ···················· | ···················· |
| W | 6160 hCoV-19/USA/AZ-TG408206/2020\|EPI_ISL_694942\|2020-0 | ···················· | ·A·················· | ····················· | ···················· | ···················· | ···················· |
| W | 6161 hCoV-19/USA/AZ-TG408210/2020\|EPI_ISL_694946\|2020-0 | ···················· | ···················· | ····················· | ···················· | ···················· | ···················· |
| W | 6162 hCoV-19/USA/AZ-TG408211/2020\|EPI_ISL_694947\|2020-0 | ···················· | ···················· | ····················· | ···················· | ···················· | ···················· |
| W | 6163 hCoV-19/USA/AZ-TG408212/2020\|EPI_ISL_694948\|2020-0 | ···················· | ·A·················· | ····················· | ···················· | ···················· | ···················· |
| W | 6164 hCoV-19/USA/AZ-TG408213/2020\|EPI_ISL_694949\|2020-0 | ···················· | ·A·················· | ····················· | ···················· | ···················· | ···················· |
| W | 6165 hCoV-19/USA/AZ-TG408305/2020\|EPI_ISL_694951\|2020-0 | ···················· | ···················· | ····················· | ···················· | ···················· | ···················· |
| W | 6166 hCoV-19/USA/AZ-TG409533/2020\|EPI_ISL_694956\|2020-0 | ···················· | ···················· | ····················· | ···················· | ···················· | ···················· |
| W | 6167 hCoV-19/USA/AZ-TG409929/2020\|EPI_ISL_694960\|2020-0 | ···················· | ·A·················· | ····················· | ···················· | ···················· | ···················· |
| W | 6168 hCoV-19/USA/AZ-TG410172/2020\|EPI_ISL_694962\|2020-0 | ···················· | ···················· | ····················· | ···················· | ···················· | ···················· |
| W | 6169 hCoV-19/USA/AZ-TG410396/2020\|EPI_ISL_694963\|2020-0 | ···················· | ·A·················· | ····················· | ···················· | ···················· | ···················· |
| W | 6170 hCoV-19/USA/AZ-TG410477/2020\|EPI_ISL_694964\|2020-0 | ···················· | ·A·················· | ····················· | ···················· | ···················· | ···················· |
| W | 6171 hCoV-19/USA/AZ-TG410650/2020\|EPI_ISL_694965\|2020-0 | ···················· | ···················· | ····················· | ···················· | ···················· | ···················· |
| W | 6172 hCoV-19/USA/AZ-TG410653/2020\|EPI_ISL_694966\|2020-0 | ···················· | ···················· | ····················· | ···················· | ···················· | ···················· |
| W | 6173 hCoV-19/USA/AZ-TG411326/2020\|EPI_ISL_694973\|2020-0 | ···················· | ···················· | ····················· | ···················· | ···················· | ···················· |
| W | 6174 hCoV-19/USA/AZ-TG411436/2020\|EPI_ISL_694974\|2020-0 | ···················· | ···················· | ····················· | ···················· | ···················· | ···················· |
| W | 6175 hCoV-19/USA/AZ-TG411558/2020\|EPI_ISL_694975\|2020-0 | ···················· | ·N·················· | ····················· | ···················· | ···················· | ···················· |
| W | 6176 hCoV-19/USA/AZ-TG411559/2020\|EPI_ISL_694976\|2020-0 | ···················· | ···················· | ····················· | ···················· | ···················· | ···················· |
| W | 6177 hCoV-19/USA/AZ-TG411811/2020\|EPI_ISL_694978\|2020-0 | ···················· | ···················· | ····················· | ···················· | ···················· | ···················· |
| W | 6178 hCoV-19/USA/AZ-TG412162/2020\|EPI_ISL_694980\|2020-0 | ···················· | ···················· | ····················· | ···················· | ···················· | ···················· |
| W | 6179 hCoV-19/USA/AZ-TG412164/2020\|EPI_ISL_694981\|2020-0 | ···················· | ···················· | ····················· | ···················· | ···················· | ···················· |
| W | 6180 hCoV-19/USA/AZ-TG412165/2020\|EPI_ISL_694982\|2020-0 | ···················· | ···················· | ····················· | ···················· | ···················· | ···················· |
| W | 6181 hCoV-19/USA/AZ-TG412167/2020\|EPI_ISL_694983\|2020-0 | ···················· | ·A·················· | ····················· | ···················· | ···················· | ···················· |
| W | 6182 hCoV-19/USA/AZ-TG412678/2020\|EPI_ISL_694984\|2020-0 | ···················· | ···················· | ····················· | ···················· | ···················· | ···················· |
| W | 6183 hCoV-19/USA/AZ-TG426799/2020\|EPI_ISL_694989\|2020-0 | ···················· | ·A·················· | ····················· | ···················· | ···················· | ···················· |

| | | | | | | | |
|---|---|---|---|---|---|---|---|
| W | 6184 hCoV-19/USA/AZ-TG442565/2020\|EPI_ISL_694994\|2020-0 | ···················· | ·A·················· | ···················· | ···················· | ···················· | ···················· |
| W | 6185 hCoV-19/USA/AZ-TG442879/2020\|EPI_ISL_694995\|2020-0 | ···················· | ·A·················· | ···················· | ···················· | ···················· | ···················· |
| W | 6186 hCoV-19/USA/AZ-TG443891/2020\|EPI_ISL_694997\|2020-0 | ···················· | ·A·················· | ···················· | ···················· | ···················· | ···················· |
| W | 6187 hCoV-19/USA/AZ-TG445156/2020\|EPI_ISL_694999\|2020-0 | ···················· | ···················· | ···················· | ···················· | ···················· | ···················· |
| W | 6188 hCoV-19/United Arab Emirates/0255/2020\|EPI_ISL_698 | ···················· | ···················· | ···················· | ···················· | ···················· | ···················· |
| W | 6189 hCoV-19/United Arab Emirates/0256/2020\|EPI_ISL_698 | ···················· | ···················· | ···················· | ···················· | ···················· | ···················· |
| W | 6190 hCoV-19/United Arab Emirates/0257/2020\|EPI_ISL_698 | ···················· | ···················· | ···················· | ···················· | ···················· | ···················· |
| W | 6191 hCoV-19/United Arab Emirates/0258/2020\|EPI_ISL_698 | ···················· | ···················· | ···················· | ···················· | ···················· | ···················· |
| W | 6192 hCoV-19/United Arab Emirates/0262/2020\|EPI_ISL_698 | ···················· | ···················· | ···················· | ···················· | ···················· | ···················· |
| W | 6193 hCoV-19/United Arab Emirates/0263/2020\|EPI_ISL_698 | ···················· | ···················· | ···················· | ···················· | ···················· | ···················· |
| W | 6194 hCoV-19/United Arab Emirates/0337/2020\|EPI_ISL_698 | ···················· | ···················· | ···················· | ···················· | ···················· | ···················· |
| W | 6195 hCoV-19/United Arab Emirates/0340/2020\|EPI_ISL_698 | ···················· | ···················· | ···················· | ···················· | ···················· | ···················· |
| W | 6196 hCoV-19/United Arab Emirates/0341/2020\|EPI_ISL_698 | ···················· | ···················· | ···················· | ···················· | ···················· | ···················· |
| W | 6197 hCoV-19/United Arab Emirates/0345/2020\|EPI_ISL_698 | ···················· | ···················· | ···················· | ···················· | ···················· | ···················· |
| W | 6198 hCoV-19/United Arab Emirates/0346/2020\|EPI_ISL_698 | ···················· | ···················· | ···················· | ···················· | ···················· | ···················· |
| W | 6199 hCoV-19/United Arab Emirates/0347/2020\|EPI_ISL_698 | ···················· | ···················· | ···················· | ···················· | ···················· | ···················· |
| W | 6200 hCoV-19/United Arab Emirates/0348/2020\|EPI_ISL_698 | ···················· | ···················· | ···················· | ···················· | ···················· | ···················· |
| W | 6201 hCoV-19/United Arab Emirates/0349/2020\|EPI_ISL_698 | ···················· | ···················· | ···················· | ···················· | ···················· | ···················· |
| W | 6202 hCoV-19/United Arab Emirates/0350/2020\|EPI_ISL_698 | ···················· | ···················· | ···················· | ···················· | ···················· | ···················· |
| W | 6203 hCoV-19/United Arab Emirates/0351/2020\|EPI_ISL_698 | ···················· | ···················· | ···················· | ···················· | ···················· | ···················· |
| W | 6204 hCoV-19/United Arab Emirates/0352/2020\|EPI_ISL_698 | ···················· | ···················· | ···················· | ···················· | ···················· | ···················· |
| W | 6205 hCoV-19/United Arab Emirates/0353/2020\|EPI_ISL_698 | ···················· | ···················· | ···················· | ···················· | ···················· | ···················· |
| W | 6206 hCoV-19/United Arab Emirates/0354/2020\|EPI_ISL_698 | ···················· | ···················· | ···················· | ···················· | ···················· | ···················· |
| W | 6207 hCoV-19/United Arab Emirates/0355/2020\|EPI_ISL_698 | ···················· | ···················· | ···················· | ···················· | ···················· | ···················· |
| W | 6208 hCoV-19/United Arab Emirates/0356/2020\|EPI_ISL_698 | ···················· | ···················· | ···················· | ···················· | ···················· | ···················· |
| W | 6209 hCoV-19/United Arab Emirates/0357/2020\|EPI_ISL_698 | ···················· | ···················· | ···················· | ···················· | ···················· | ···················· |
| W | 6210 hCoV-19/United Arab Emirates/0358/2020\|EPI_ISL_698 | ···················· | ···················· | ···················· | ···················· | ···················· | ···················· |
| W | 6211 hCoV-19/United Arab Emirates/0359/2020\|EPI_ISL_698 | ···················· | ···················· | ···················· | ···················· | ···················· | ···················· |
| W | 6212 hCoV-19/United Arab Emirates/0360/2020\|EPI_ISL_698 | ···················· | ···················· | ···················· | ···················· | ···················· | ···················· |
| W | 6213 hCoV-19/United Arab Emirates/0361/2020\|EPI_ISL_698 | ···················· | ···················· | ···················· | ···················· | ···················· | ···················· |
| W | 6214 hCoV-19/United Arab Emirates/0362/2020\|EPI_ISL_698 | ···················· | ···················· | ···················· | ···················· | ···················· | ···················· |
| W | 6215 hCoV-19/United Arab Emirates/0363/2020\|EPI_ISL_698 | ···················· | ···················· | ···················· | ···················· | ···················· | ···················· |
| W | 6216 hCoV-19/United Arab Emirates/0364/2020\|EPI_ISL_698 | ···················· | ···················· | ···················· | ···················· | ···················· | ···················· |
| W | 6217 hCoV-19/United Arab Emirates/0365/2020\|EPI_ISL_698 | ···················· | ···················· | ···················· | ···················· | ···················· | ···················· |
| W | 6218 hCoV-19/United Arab Emirates/0366/2020\|EPI_ISL_698 | ···················· | ···················· | ···················· | ···················· | ···················· | ···················· |
| W | 6219 hCoV-19/United Arab Emirates/0367/2020\|EPI_ISL_698 | ···················· | ···················· | ···················· | ···················· | ···················· | ···················· |
| W | 6220 hCoV-19/United Arab Emirates/0368/2020\|EPI_ISL_698 | ···················· | ···················· | ···················· | ···················· | ···················· | ···················· |
| W | 6221 hCoV-19/United Arab Emirates/0369/2020\|EPI_ISL_698 | ···················· | ···················· | ···················· | ···················· | ···················· | ···················· |
| W | 6222 hCoV-19/United Arab Emirates/0370/2020\|EPI_ISL_698 | ···················· | ···················· | ···················· | ···················· | ···················· | ···················· |
| W | 6223 hCoV-19/United Arab Emirates/0371/2020\|EPI_ISL_698 | ···················· | ···················· | ···················· | ···················· | ···················· | ···················· |
| W | 6224 hCoV-19/United Arab Emirates/0372/2020\|EPI_ISL_698 | ···················· | ···················· | ···················· | ···················· | ···················· | ···················· |
| W | 6225 hCoV-19/United Arab Emirates/0373/2020\|EPI_ISL_698 | ···················· | ···················· | ···················· | ···················· | ···················· | ···················· |
| W | 6226 hCoV-19/United Arab Emirates/0374/2020\|EPI_ISL_698 | ···················· | ···················· | ···················· | ···················· | ···················· | ···················· |
| W | 6227 hCoV-19/United Arab Emirates/0375/2020\|EPI_ISL_698 | ···················· | ···················· | ···················· | ···················· | ···················· | ···················· |
| W | 6228 hCoV-19/United Arab Emirates/0376/2020\|EPI_ISL_698 | ···················· | ···················· | ···················· | ···················· | ···················· | ···················· |
| W | 6229 hCoV-19/United Arab Emirates/0377/2020\|EPI_ISL_698 | ···················· | ···················· | ···················· | ···················· | ···················· | ···················· |
| W | 6230 hCoV-19/United Arab Emirates/0378/2020\|EPI_ISL_698 | ···················· | ···················· | ···················· | ···················· | ···················· | ···················· |
| W | 6231 hCoV-19/United Arab Emirates/0379/2020\|EPI_ISL_698 | ···················· | ···················· | ···················· | ···················· | ···················· | ···················· |
| W | 6232 hCoV-19/United Arab Emirates/0380/2020\|EPI_ISL_698 | ···················· | ···················· | ···················· | ···················· | ···················· | ···················· |
| W | 6233 hCoV-19/United Arab Emirates/0381/2020\|EPI_ISL_698 | ···················· | ···················· | ···················· | ···················· | ···················· | ···················· |
| W | 6234 hCoV-19/India/MH-ACTREC-112/2020\|EPI_ISL_699768\|20 | ···················· | ···················· | ···················· | ···················· | ···················· | ···················· |
| W | 6235 hCoV-19/India/MH-ACTREC-113/2020\|EPI_ISL_699769\|20 | ···················· | ···················· | ···················· | ···················· | ···················· | ···················· |
| W | 6236 hCoV-19/India/MH-ACTREC-120/2020\|EPI_ISL_699776\|20 | ···················· | ···················· | ···················· | ···················· | ···················· | ···················· |
| W | 6237 hCoV-19/India/MH-ACTREC-121/2020\|EPI_ISL_699777\|20 | ···················· | ···················· | ···················· | ···················· | ···················· | ···················· |
| W | 6240 hCoV-19/India/MH-ACTREC-193/2020\|EPI_ISL_699847\|20 | ···················· | ···················· | ···················· | ···················· | ···················· | ···················· |
| W | 6243 hCoV-19/India/MH-ACTREC-198/2020\|EPI_ISL_699852\|20 | ···················· | ···················· | ···················· | ···················· | ···················· | ···················· |
| W | 6244 hCoV-19/India/MH-ACTREC-199/2020\|EPI_ISL_699853\|20 | ···················· | ···················· | ···················· | ···················· | ···················· | ···················· |
| W | 6245 hCoV-19/India/MH-ACTREC-200/2020\|EPI_ISL_699854\|20 | ···················· | ···················· | ···················· | ···················· | ···················· | ···················· |
| W | 6247 hCoV-19/India/MH-ACTREC-202/2020\|EPI_ISL_699856\|20 | ···················· | ···················· | ···················· | ···················· | ···················· | ···················· |
| W | 6248 hCoV-19/India/MH-ACTREC-203/2020\|EPI_ISL_699857\|20 | ···················· | ···················· | ···················· | ···················· | ···················· | ···················· |
| W | 6249 hCoV-19/India/MH-ACTREC-204/2020\|EPI_ISL_699858\|20 | ···················· | ···················· | ···················· | ···················· | ···················· | ···················· |
| W | 6250 hCoV-19/India/MH-ACTREC-205/2020\|EPI_ISL_699859\|20 | ···················· | ···················· | ···················· | ···················· | ···················· | ···················· |
| W | 6251 hCoV-19/India/MH-ACTREC-206/2020\|EPI_ISL_699860\|20 | ···················· | ···················· | ···················· | ···················· | ···················· | ···················· |
| W | 6252 hCoV-19/India/MH-ACTREC-207/2020\|EPI_ISL_699861\|20 | ···················· | ···················· | ···················· | ···················· | ···················· | ···················· |
| W | 6253 hCoV-19/India/MH-ACTREC-208/2020\|EPI_ISL_699862\|20 | ···················· | ···················· | ···················· | ···················· | ···················· | ···················· |
| W | 6255 hCoV-19/India/MH-ACTREC-210/2020\|EPI_ISL_699864\|20 | ···················· | ···················· | ···················· | ···················· | ···················· | ···················· |
| W | 6256 hCoV-19/India/MH-ACTREC-211/2020\|EPI_ISL_699865\|20 | ···················· | ···················· | ···················· | ···················· | ···················· | ···················· |
| W | 6257 hCoV-19/India/MH-ACTREC-212/2020\|EPI_ISL_699866\|20 | ···················· | ···················· | ···················· | ···················· | ···················· | ···················· |
| W | 6258 hCoV-19/India/MH-ACTREC-213/2020\|EPI_ISL_699867\|20 | ···················· | ···················· | ···················· | ···················· | ···················· | ···················· |
| W | 6259 hCoV-19/India/MH-ACTREC-214/2020\|EPI_ISL_699868\|20 | ···················· | ···················· | ···················· | ···················· | ···················· | ···················· |
| W | 6260 hCoV-19/India/MH-ACTREC-215/2020\|EPI_ISL_699869\|20 | ···················· | ···················· | ···················· | ···················· | ···················· | ···················· |
| W | 6261 hCoV-19/India/MH-ACTREC-216/2020\|EPI_ISL_699870\|20 | ···················· | ···················· | ···················· | ···················· | ···················· | ···················· |
| W | 6262 hCoV-19/India/MH-ACTREC-217/2020\|EPI_ISL_699871\|20 | ···················· | ···················· | ···················· | ···················· | ···················· | ···················· |
| W | 6263 hCoV-19/India/MH-ACTREC-218/2020\|EPI_ISL_699872\|20 | ···················· | ···················· | ···················· | ···················· | ···················· | ···················· |
| W | 6264 hCoV-19/India/MH-ACTREC-219/2020\|EPI_ISL_699873\|20 | ···················· | ···················· | ···················· | ···················· | ···················· | ···················· |
| W | 6265 hCoV-19/India/MH-ACTREC-220/2020\|EPI_ISL_699874\|20 | ···················· | ···················· | ···················· | ···················· | ···················· | ···················· |
| W | 6266 hCoV-19/India/MH-ACTREC-221/2020\|EPI_ISL_699875\|20 | ···················· | ···················· | ···················· | ···················· | ···················· | ···················· |
| W | 6267 hCoV-19/India/MH-ACTREC-222/2020\|EPI_ISL_699876\|20 | ···················· | ···················· | ···················· | ···················· | ···················· | ···················· |
| W | 6268 hCoV-19/India/MH-ACTREC-223/2020\|EPI_ISL_699877\|20 | ···················· | ···················· | ···················· | ···················· | ···················· | ···················· |
| W | 6269 hCoV-19/India/MH-ACTREC-224/2020\|EPI_ISL_699878\|20 | ···················· | ···················· | ···················· | ···················· | ···················· | ···················· |

| | | | | | | | | |
|---|---|---|---|---|---|---|---|---|
| W | 6270 | hCoV-19/India/MH-ACTREC-225/2020\|EPI_ISL_699879\|20 | ···················· | ···················· | ···················· | ···················· | ···················· | ···················· |
| W | 6271 | hCoV-19/India/MH-ACTREC-226/2020\|EPI_ISL_699880\|20 | ···················· | ···················· | ···················· | ···················· | ···················· | ···················· |
| W | 6272 | hCoV-19/India/MH-ACTREC-227/2020\|EPI_ISL_699881\|20 | ···················· | ···················· | ···················· | ···················· | ···················· | ···················· |
| W | 6273 | hCoV-19/India/MH-ACTREC-228/2020\|EPI_ISL_699882\|20 | ···················· | ···················· | ···················· | ···················· | ···················· | ···················· |
| W | 6274 | hCoV-19/India/MH-ACTREC-230/2020\|EPI_ISL_699883\|20 | ···················· | ···················· | ···················· | ···················· | ···················· | ···················· |
| W | 6275 | hCoV-19/India/MH-ACTREC-231/2020\|EPI_ISL_699884\|20 | ···················· | ···················· | ···················· | ···················· | ···················· | ···················· |
| W | 6276 | hCoV-19/India/MH-ACTREC-232/2020\|EPI_ISL_699885\|20 | ···················· | ···················· | ···················· | ···················· | ···················· | ···················· |
| W | 6277 | hCoV-19/India/MH-ACTREC-233/2020\|EPI_ISL_699886\|20 | ···················· | ···················· | ···················· | ···················· | ···················· | ···················· |
| W | 6278 | hCoV-19/India/MH-ACTREC-234/2020\|EPI_ISL_699887\|20 | ···················· | ···················· | ···················· | ···················· | ···················· | ···················· |
| W | 6279 | hCoV-19/USA/TX-DSHS-1615/2020\|EPI_ISL_700739\|2020- | ···················· | ···················· | ···················· | ···················· | ···················· | ···················· |
| W | 6280 | hCoV-19/USA/TX-DSHS-1595/2020\|EPI_ISL_700740\|2020- | ···················· | ···················· | ···················· | ···················· | ···················· | ···················· |
| W | 6281 | hCoV-19/USA/TX-DSHS-1606/2020\|EPI_ISL_700741\|2020- | ···················· | ···················· | ···················· | ···················· | ···················· | ···················· |
| W | 6282 | hCoV-19/England/QEUH-BCCDD1/2020\|EPI_ISL_704272\|20 | ···················· | ···················· | ···················· | ···················· | ···················· | ···················· |
| W | 6283 | hCoV-19/England/QEUH-BCE276/2020\|EPI_ISL_704274\|20 | ···················· | ···················· | ···················· | ···················· | ···················· | ···················· |
| W | 6284 | hCoV-19/England/QEUH-BCB561/2020\|EPI_ISL_704275\|20 | ···················· | ···················· | ···················· | ···················· | ···················· | ···················· |
| W | 6285 | hCoV-19/England/QEUH-BCCE1A/2020\|EPI_ISL_704278\|20 | ···················· | ···················· | ···················· | ···················· | ···················· | ···················· |
| W | 6286 | hCoV-19/England/QEUH-BCB5AD/2020\|EPI_ISL_704279\|20 | ···················· | ·A·················· | ···················· | ···················· | ···················· | ···················· |
| W | 6287 | hCoV-19/England/QEUH-BCD954/2020\|EPI_ISL_704281\|20 | ···················· | ···················· | ···················· | ···················· | ···················· | ···················· |
| W | 6288 | hCoV-19/England/QEUH-BCE4CB/2020\|EPI_ISL_704284\|20 | ···················· | ···················· | ···················· | ···················· | ···················· | ···················· |
| W | 6289 | hCoV-19/England/QEUH-BCBB23/2020\|EPI_ISL_704287\|20 | ···················· | ···················· | ···················· | ···················· | ···················· | ···················· |
| W | 6290 | hCoV-19/England/QEUH-BCE944/2020\|EPI_ISL_704288\|20 | ···················· | ···················· | ···················· | ···················· | ···················· | ···················· |
| W | 6291 | hCoV-19/England/QEUH-BCCDA4/2020\|EPI_ISL_704293\|20 | ···················· | ···················· | ···················· | ···················· | ···················· | ···················· |
| W | 6292 | hCoV-19/England/QEUH-BCD3B0/2020\|EPI_ISL_704295\|20 | ···················· | ···················· | ···················· | ···················· | ······T············· | ···················· |
| W | 6293 | hCoV-19/England/QEUH-BCE20D/2020\|EPI_ISL_704296\|20 | ···················· | ···················· | ···················· | ···················· | ···················· | ···················· |
| W | 6294 | hCoV-19/England/QEUH-BCE58C/2020\|EPI_ISL_704298\|20 | ···················· | ···················· | ···················· | ···················· | ···················· | ···················· |
| W | 6295 | hCoV-19/England/CAMC-BBDC9A/2020\|EPI_ISL_705619\|20 | ···················· | ···················· | ···················· | ···················· | ···················· | ···················· |
| W | 6296 | hCoV-19/Scotland/QEUH-A35EE0/2020\|EPI_ISL_706900\|2 | ···················· | ···················· | ···················· | ···················· | ···················· | ···················· |
| W | 6297 | hCoV-19/Scotland/QEUH-A35EFF/2020\|EPI_ISL_706901\|2 | ···················· | ···················· | ···················· | ···················· | ···················· | ···················· |
| W | 6298 | hCoV-19/England/QEUH-A35F0B/2020\|EPI_ISL_706902\|20 | ···················· | ···················· | ···················· | ···················· | ···················· | ···················· |
| W | 6299 | hCoV-19/England/QEUH-A3619B/2020\|EPI_ISL_706903\|20 | ···················· | ···················· | ···················· | ···················· | ···················· | ···················· |
| W | 6300 | hCoV-19/Wales/QEUH-A361B9/2020\|EPI_ISL_706904\|2020 | ···················· | ···················· | ···················· | ···················· | ···················· | ···················· |
| W | 6301 | hCoV-19/England/QEUH-A36465/2020\|EPI_ISL_706905\|20 | ···················· | ···················· | ···················· | ···················· | ···················· | ···················· |
| W | 6302 | hCoV-19/England/QEUH-A36474/2020\|EPI_ISL_706906\|20 | ···················· | ···················· | ···················· | ···················· | ···················· | ···················· |
| W | 6303 | hCoV-19/England/QEUH-A36483/2020\|EPI_ISL_706907\|20 | ···················· | ···················· | ···················· | ···················· | ···················· | ···················· |
| W | 6304 | hCoV-19/England/QEUH-A36492/2020\|EPI_ISL_706908\|20 | ···················· | ···················· | ···················· | ···················· | ···················· | ···················· |
| W | 6305 | hCoV-19/England/QEUH-A364A1/2020\|EPI_ISL_706909\|20 | ···················· | ···················· | ···················· | ···················· | ···················· | ···················· |
| W | 6306 | hCoV-19/England/QEUH-A364B0/2020\|EPI_ISL_706910\|20 | ···················· | ···················· | ···················· | ···················· | ···················· | ···················· |
| W | 6307 | hCoV-19/England/QEUH-A36517/2020\|EPI_ISL_706911\|20 | ···················· | ···················· | ···················· | ···················· | ···················· | ···················· |
| W | 6308 | hCoV-19/England/QEUH-A36544/2020\|EPI_ISL_706912\|20 | ···················· | ···················· | ···················· | ···················· | ···················· | ···················· |
| W | 6309 | hCoV-19/England/QEUH-A366BA/2020\|EPI_ISL_706913\|20 | ···················· | ···················· | ···················· | ···················· | ······T············· | ···················· |
| W | 6310 | hCoV-19/England/QEUH-A366D8/2020\|EPI_ISL_706914\|20 | ···················· | ···················· | ···················· | ···················· | ···················· | ···················· |
| W | 6311 | hCoV-19/England/QEUH-A366E7/2020\|EPI_ISL_706915\|20 | ···················· | ···················· | ···················· | ···················· | ···················· | ···················· |
| W | 6312 | hCoV-19/England/QEUH-A3676C/2020\|EPI_ISL_706916\|20 | ···················· | ···················· | ···················· | ···················· | ···················· | ···················· |
| W | 6313 | hCoV-19/England/QEUH-A3677B/2020\|EPI_ISL_706917\|20 | ···················· | ···················· | ···················· | ···················· | ···················· | ···················· |
| W | 6314 | hCoV-19/England/QEUH-A367A8/2020\|EPI_ISL_706918\|20 | ···················· | ···················· | ···················· | ···················· | ···················· | ···················· |
| W | 6315 | hCoV-19/England/QEUH-A367B7/2020\|EPI_ISL_706919\|20 | ···················· | ···················· | ···················· | ···················· | ···················· | ···················· |
| W | 6316 | hCoV-19/England/QEUH-A36878/2020\|EPI_ISL_706920\|20 | ···················· | ···················· | ···················· | ···················· | ···················· | ···················· |
| W | 6317 | hCoV-19/England/QEUH-A36887/2020\|EPI_ISL_706921\|20 | ···················· | ···················· | ···················· | ···················· | ···················· | ···················· |
| W | 6318 | hCoV-19/England/QEUH-A36896/2020\|EPI_ISL_706922\|20 | ···················· | ···················· | ···················· | ···················· | ···················· | ···················· |
| W | 6319 | hCoV-19/England/QEUH-A368B4/2020\|EPI_ISL_706923\|20 | ···················· | ···················· | ···················· | ···················· | ···················· | ···················· |
| W | 6320 | hCoV-19/England/QEUH-A36C21/2020\|EPI_ISL_706924\|20 | ···················· | ···················· | ···················· | ···················· | ···················· | ···················· |
| W | 6321 | hCoV-19/Scotland/QEUH-A36C30/2020\|EPI_ISL_706925\|2 | ···················· | ···················· | ···················· | ···················· | ···················· | ···················· |
| W | 6322 | hCoV-19/Scotland/QEUH-A36C4F/2020\|EPI_ISL_706926\|2 | ···················· | ···················· | ···················· | ···················· | ···················· | ···················· |
| W | 6323 | hCoV-19/England/QEUH-A36C8B/2020\|EPI_ISL_706927\|20 | ···················· | ···················· | ···················· | ···················· | ···················· | ···················· |
| W | 6324 | hCoV-19/England/CAMC-BFB0D0/2020\|EPI_ISL_709100\|20 | ···················· | ···················· | ···················· | ···················· | ···················· | ···················· |
| W | 6325 | hCoV-19/England/CAMC-BFB3F5/2020\|EPI_ISL_709101\|20 | ···················· | ···················· | ···················· | ···················· | ···················· | ···················· |
| W | 6327 | hCoV-19/England/QEUH-BF0D6B/2020\|EPI_ISL_709108\|20 | ···················· | ···················· | ···················· | ···················· | ···················· | ···················· |
| W | 6328 | hCoV-19/England/CAMC-BFBC18/2020\|EPI_ISL_709116\|20 | ···················· | ···················· | ···················· | ···················· | ···················· | ···················· |
| W | 6329 | hCoV-19/England/CAMC-BFB9A8/2020\|EPI_ISL_709117\|20 | ···················· | ···················· | ···················· | ···················· | ······T············· | ···················· |
| W | 6330 | hCoV-19/England/CAMC-BFBD7F/2020\|EPI_ISL_709118\|20 | ···················· | ···················· | ···················· | ···················· | ···················· | ···················· |
| W | 6331 | hCoV-19/England/QEUH-BF0A46/2020\|EPI_ISL_709119\|20 | ···················· | ···················· | ···················· | ···················· | ···················· | ···················· |
| W | 6332 | hCoV-19/England/CAMC-BFB647/2020\|EPI_ISL_709122\|20 | ···················· | ···················· | ···················· | ···················· | ······T············· | ···················· |
| W | 6333 | hCoV-19/England/CAMC-BFC02A/2020\|EPI_ISL_709123\|20 | ···················· | ···················· | ···················· | ···················· | ···················· | ···················· |
| W | 6334 | hCoV-19/England/CAMC-BFB9B7/2020\|EPI_ISL_709124\|20 | ···················· | ···················· | ···················· | ···················· | ···················· | ···················· |
| W | 6335 | hCoV-19/England/CAMC-BFBA78/2020\|EPI_ISL_709130\|20 | ···················· | ···················· | ···················· | ···················· | ······T············· | ···················· |
| W | 6336 | hCoV-19/England/QEUH-BF0BDA/2020\|EPI_ISL_709131\|20 | ···················· | ···················· | ···················· | ···················· | ···················· | ···················· |
| W | 6337 | hCoV-19/England/QEUH-BF0985/2020\|EPI_ISL_709132\|20 | ···················· | ···················· | ···················· | ···················· | ···················· | ···················· |
| W | 6338 | hCoV-19/England/QEUH-BF0B52/2020\|EPI_ISL_709133\|20 | ···················· | ···················· | ···················· | ···················· | ···················· | ···················· |
| W | 6339 | hCoV-19/England/QEUH-BF119B/2020\|EPI_ISL_709135\|20 | ···················· | ···················· | ···················· | ···················· | ···················· | ···················· |
| W | 6340 | hCoV-19/England/QEUH-BF118C/2020\|EPI_ISL_709139\|20 | ···················· | ···················· | ···················· | ···················· | ···················· | ···················· |
| W | 6341 | hCoV-19/England/QEUH-BF0ABF/2020\|EPI_ISL_709140\|20 | ···················· | ···················· | ···················· | ···················· | ···················· | ···················· |
| W | 6342 | hCoV-19/England/QEUH-BF1140/2020\|EPI_ISL_709141\|20 | ···················· | ···················· | ···················· | ···················· | ···················· | ···················· |
| W | 6343 | hCoV-19/England/QEUH-BF0B43/2020\|EPI_ISL_709142\|20 | ···················· | ···················· | ···················· | ···················· | ···················· | ···················· |
| W | 6344 | hCoV-19/England/CAMC-BFB665/2020\|EPI_ISL_709143\|20 | ···················· | ···················· | ···················· | ···················· | ···················· | ···················· |
| W | 6345 | hCoV-19/England/QEUH-BF0C7D/2020\|EPI_ISL_709144\|20 | ···················· | ···················· | ···················· | ···················· | ···················· | ···················· |
| W | 6346 | hCoV-19/England/QEUH-BF0B61/2020\|EPI_ISL_709150\|20 | ···················· | ···················· | ···················· | ···················· | ···················· | ···················· |
| W | 6347 | hCoV-19/England/QEUH-BF0ACE/2020\|EPI_ISL_709151\|20 | ···················· | ···················· | ···················· | ···················· | ······T············· | ···················· |
| W | 6348 | hCoV-19/England/CAMC-BFB2CB/2020\|EPI_ISL_709152\|20 | ···················· | ···················· | ···················· | ···················· | ···················· | ···················· |
| W | 6349 | hCoV-19/England/CAMC-BFB01C/2020\|EPI_ISL_709153\|20 | ···················· | ···················· | ···················· | ···················· | ···················· | ···················· |
| W | 6350 | hCoV-19/England/CAMC-BFB155/2020\|EPI_ISL_709154\|20 | ···················· | ···················· | ···················· | ···················· | ···················· | ···················· |

| | | | | | | | | |
|---|---|---|---|---|---|---|---|---|
| W | 6351 | hCoV-19/England/QEUH-BF0967/2020\|EPI_ISL_709159\|20 | ···················· | ···················· | ····················· | ···················· | ···················· | ···················· |
| W | 6352 | hCoV-19/England/QEUH-BF0D7A/2020\|EPI_ISL_709160\|20 | ···················· | ···················· | ····················· | ···················· | ···················· | ···················· |
| W | 6353 | hCoV-19/England/QEUH-BF0B16/2020\|EPI_ISL_709161\|20 | ···················· | ···················· | ····················· | ···················· | ···················· | ···················· |
| W | 6354 | hCoV-19/England/CAMC-BFB999/2020\|EPI_ISL_709166\|20 | ···················· | ···················· | ····················· | ···················· | ···················· | ···················· |
| W | 6355 | hCoV-19/England/CAMC-BFBA1E/2020\|EPI_ISL_709167\|20 | ···················· | ···················· | ····················· | ···················· | ····T··············· | ···················· |
| W | 6356 | hCoV-19/England/CAMC-BFBFF1/2020\|EPI_ISL_709169\|20 | ···················· | ···················· | ····················· | ···················· | ···················· | ···················· |
| W | 6357 | hCoV-19/England/QEUH-BF0C9B/2020\|EPI_ISL_709170\|20 | ···················· | ···················· | ····················· | ···················· | ······T············· | ···················· |
| W | 6358 | hCoV-19/England/QEUH-BF19EE/2020\|EPI_ISL_709171\|20 | ···················· | ···················· | ····················· | ···················· | ···················· | ···················· |
| W | 6359 | hCoV-19/England/QEUH-BF0B07/2020\|EPI_ISL_709174\|20 | ···················· | ···················· | ····················· | ···················· | ···················· | ···················· |
| W | 6360 | hCoV-19/England/CAMC-BFBD15/2020\|EPI_ISL_709177\|20 | ···················· | ···················· | ····················· | ···················· | ······T············· | ···················· |
| W | 6361 | hCoV-19/England/CAMC-BFBCEB/2020\|EPI_ISL_709178\|20 | ···················· | ···················· | ····················· | ···················· | ···················· | ···················· |
| W | 6362 | hCoV-19/England/CAMC-BFBE8B/2020\|EPI_ISL_709180\|20 | ···················· | ···················· | ····················· | ···················· | ···················· | ···················· |
| W | 6363 | hCoV-19/England/QEUH-BF0D2F/2020\|EPI_ISL_709189\|20 | ···················· | ···················· | ····················· | ···················· | ···················· | ···················· |
| W | 6364 | hCoV-19/England/QEUH-BF0B8F/2020\|EPI_ISL_709190\|20 | ···················· | ···················· | ····················· | ···················· | ···················· | ···················· |
| W | 6365 | hCoV-19/England/QEUH-BF0958/2020\|EPI_ISL_709191\|20 | ···················· | ···················· | ····················· | ···················· | ···················· | ···················· |
| W | 6366 | hCoV-19/England/CAMC-BFC084/2020\|EPI_ISL_709193\|20 | ···················· | ···················· | ····················· | ···················· | ···················· | ···················· |
| W | 6367 | hCoV-19/England/CAMC-BFB46B/2020\|EPI_ISL_709196\|20 | ···················· | ···················· | ····················· | ···················· | ···················· | ···················· |
| W | 6368 | hCoV-19/England/QEUH-BF0994/2020\|EPI_ISL_709199\|20 | ···················· | ···················· | ····················· | ···················· | ···················· | ···················· |
| W | 6369 | hCoV-19/USA/CA-LACPHL-AA00002/2020\|EPI_ISL_710100\| | ···················· | ···················· | ····················· | ···················· | ···················· | ···················· |
| W | 6370 | hCoV-19/USA/CA-LACPHL-AA00003/2020\|EPI_ISL_710101\| | ···················· | ···················· | ····················· | ···················· | ···················· | ···················· |
| W | 6371 | hCoV-19/USA/CA-LACPHL-AA00004/2020\|EPI_ISL_710102\| | ···················· | ···················· | ····················· | ···················· | ···················· | ···················· |
| W | 6372 | hCoV-19/USA/CA-LACPHL-AA00005/2020\|EPI_ISL_710103\| | ···················· | ···················· | ····················· | ···················· | ···················· | ···················· |
| W | 6373 | hCoV-19/USA/CA-LACPHL-AA00006/2020\|EPI_ISL_710104\| | ···················· | ·A·················· | ····················· | ···················· | ···················· | ···················· |
| W | 6374 | hCoV-19/USA/CA-LACPHL-AA00008/2020\|EPI_ISL_710106\| | ···················· | ···················· | ····················· | ···················· | ···················· | ···················· |
| W | 6375 | hCoV-19/USA/CA-LACPHL-AA00009/2020\|EPI_ISL_710107\| | ···················· | ·A·················· | ····················· | ···················· | ···················· | ···················· |
| W | 6376 | hCoV-19/USA/CA-LACPHL-AA00010/2020\|EPI_ISL_710108\| | ···················· | ·A·················· | ····················· | ···················· | ···················· | ···················· |
| W | 6377 | hCoV-19/USA/CA-LACPHL-AA00011/2020\|EPI_ISL_710109\| | ········T··········· | ···················· | ····················· | ···················· | ···················· | ···················· |
| W | 6378 | hCoV-19/USA/CA-LACPHL-AA00014/2020\|EPI_ISL_710112\| | ···················· | ···················· | ····················· | ···················· | ···················· | ···················· |
| W | 6379 | hCoV-19/USA/CA-LACPHL-AA00015/2020\|EPI_ISL_710113\| | ···················· | ·A·················· | ····················· | ···················· | ···················· | ···················· |
| W | 6380 | hCoV-19/USA/CA-LACPHL-AA00018/2020\|EPI_ISL_710116\| | ···················· | ···················· | ····················· | ···················· | ···················· | ···················· |
| W | 6381 | hCoV-19/Australia/NSW1221/2020\|EPI_ISL_710120\|2020 | ···················· | ·A·················· | ····················· | ···················· | ···················· | ···A················ |
| W | 6382 | hCoV-19/Australia/NSW1223/2020\|EPI_ISL_710121\|2020 | ···················· | ···················· | ····················· | ···················· | ···················· | ···················· |
| W | 6383 | hCoV-19/Australia/NSW1224/2020\|EPI_ISL_710122\|2020 | ···················· | ···················· | ····················· | ···················· | ···················· | ···················· |
| W | 6384 | hCoV-19/Australia/NSW1225/2020\|EPI_ISL_710123\|2020 | ···················· | ·A·················· | ····················· | ···················· | ···················· | ···················· |
| W | 6385 | hCoV-19/Australia/NSW1226/2020\|EPI_ISL_710124\|2020 | ···················· | ···················· | ····················· | ···················· | ···················· | ···················· |
| W | 6386 | hCoV-19/Australia/NSW1227/2020\|EPI_ISL_710125\|2020 | ···················· | ···················· | ····················· | ···················· | ···················· | ···················· |
| W | 6387 | hCoV-19/Australia/NSW1228/2020\|EPI_ISL_710126\|2020 | ···················· | ···················· | ····················· | ···················· | ···················· | ···················· |
| W | 6388 | hCoV-19/Australia/NSW1229/2020\|EPI_ISL_710127\|2020 | ···················· | ·A·················· | ····················· | ···················· | ···················· | ···A················ |
| W | 6389 | hCoV-19/Australia/NSW1230/2020\|EPI_ISL_710128\|2020 | ···················· | ·············A······ | ····················· | ···················· | ···················· | ···················· |
| W | 6390 | hCoV-19/Australia/NSW1231/2020\|EPI_ISL_710129\|2020 | ···················· | ···················· | ····················· | ···················· | ···················· | ···················· |
| W | 6391 | hCoV-19/USA/CO-CDPHE-2004220685/2020\|EPI_ISL_71018 | ···················· | ···················· | ····················· | ···················· | ···················· | ···················· |
| W | 6394 | hCoV-19/USA/CO-CDPHE-2004220698/2020\|EPI_ISL_71018 | ···················· | ···················· | ····················· | ···················· | ··N················· | ···················· |
| W | 6396 | hCoV-19/USA/CO-CDPHE-2004220733/2020\|EPI_ISL_71018 | ···················· | ···················· | ····················· | ···················· | ···················· | ···················· |
| W | 6397 | hCoV-19/USA/CO-CDPHE-2004220748/2020\|EPI_ISL_71018 | ···················· | ···················· | ····················· | ···················· | ···················· | ···················· |
| W | 6398 | hCoV-19/USA/CO-CDPHE-2004220752/2020\|EPI_ISL_71018 | ···················· | ···················· | ····················· | ···················· | ···················· | ···················· |
| W | 6399 | hCoV-19/USA/CO-CDPHE-2004220754/2020\|EPI_ISL_71018 | ···················· | ···················· | ····················· | ···················· | ···················· | ···················· |
| W | 6400 | hCoV-19/USA/CO-CDPHE-2004220755/2020\|EPI_ISL_71018 | ···················· | ···················· | ····················· | ···················· | ·NN················· | ···················· |
| W | 6401 | hCoV-19/United Arab Emirates/0259/2020\|EPI_ISL_698 | ···················· | ···················· | ····················· | ···················· | ···················· | ···················· |
| W | 6402 | hCoV-19/United Arab Emirates/0260/2020\|EPI_ISL_698 | ···················· | ···················· | ····················· | ···················· | ···················· | ···················· |
| W | 6403 | hCoV-19/United Arab Emirates/0261/2020\|EPI_ISL_698 | ···················· | ···················· | ····················· | ···················· | ···················· | ···················· |
| W | 6404 | hCoV-19/United Arab Emirates/0264/2020\|EPI_ISL_698 | ···················· | ···················· | ····················· | ···················· | ···················· | ···················· |
| W | 6405 | hCoV-19/United Arab Emirates/0265/2020\|EPI_ISL_698 | ···················· | ···················· | ····················· | ···················· | ···················· | ···················· |
| W | 6406 | hCoV-19/United Arab Emirates/0266/2020\|EPI_ISL_698 | ···················· | ···················· | ····················· | ···················· | ···················· | ···················· |
| W | 6407 | hCoV-19/United Arab Emirates/0267/2020\|EPI_ISL_698 | ···················· | ···················· | ····················· | ···················· | ···················· | ···················· |
| W | 6408 | hCoV-19/United Arab Emirates/0268/2020\|EPI_ISL_698 | ···················· | ···················· | ····················· | ···················· | ···················· | ···················· |
| W | 6409 | hCoV-19/United Arab Emirates/0269/2020\|EPI_ISL_698 | ···················· | ···················· | ····················· | ···················· | ···················· | ···················· |
| W | 6410 | hCoV-19/United Arab Emirates/0270/2020\|EPI_ISL_698 | ···················· | ···················· | ····················· | ···················· | ···················· | ···················· |
| W | 6411 | hCoV-19/United Arab Emirates/0271/2020\|EPI_ISL_698 | ···················· | ···················· | ····················· | ···················· | ···················· | ···················· |
| W | 6412 | hCoV-19/United Arab Emirates/0272/2020\|EPI_ISL_698 | ···················· | ···················· | ····················· | ···················· | ···················· | ···················· |
| W | 6413 | hCoV-19/United Arab Emirates/0274/2020\|EPI_ISL_698 | ···················· | ···················· | ····················· | ···················· | ···················· | ···················· |
| W | 6414 | hCoV-19/United Arab Emirates/0275/2020\|EPI_ISL_698 | ···················· | ···················· | ····················· | ···················· | ···················· | ···················· |
| W | 6415 | hCoV-19/United Arab Emirates/0276/2020\|EPI_ISL_698 | ···················· | ···················· | ····················· | ···················· | ···················· | ···················· |
| W | 6416 | hCoV-19/United Arab Emirates/0277/2020\|EPI_ISL_698 | ···················· | ···················· | ····················· | ···················· | ···················· | ···················· |
| W | 6417 | hCoV-19/United Arab Emirates/0278/2020\|EPI_ISL_698 | ···················· | ···················· | ····················· | ···················· | ···················· | ···················· |
| W | 6418 | hCoV-19/United Arab Emirates/0279/2020\|EPI_ISL_698 | ···················· | ···················· | ····················· | ···················· | ···················· | ···················· |
| W | 6419 | hCoV-19/United Arab Emirates/0280/2020\|EPI_ISL_698 | ···················· | ···················· | ····················· | ···················· | ···················· | ···················· |
| W | 6420 | hCoV-19/United Arab Emirates/0281/2020\|EPI_ISL_698 | ···················· | ···················· | ····················· | ···················· | ···················· | ···················· |
| W | 6421 | hCoV-19/United Arab Emirates/0282/2020\|EPI_ISL_698 | ···················· | ···················· | ····················· | ···················· | ···················· | ···················· |
| W | 6422 | hCoV-19/United Arab Emirates/0283/2020\|EPI_ISL_698 | ···················· | ···················· | ····················· | ···················· | ···················· | ···················· |
| W | 6423 | hCoV-19/United Arab Emirates/0284/2020\|EPI_ISL_698 | ···················· | ···················· | ····················· | ···················· | ···················· | ···················· |
| W | 6424 | hCoV-19/United Arab Emirates/0285/2020\|EPI_ISL_698 | ···················· | ···················· | ····················· | ···················· | ···················· | ···················· |
| W | 6425 | hCoV-19/United Arab Emirates/0286/2020\|EPI_ISL_698 | ···················· | ···················· | ····················· | ···················· | ···················· | ···················· |
| W | 6426 | hCoV-19/United Arab Emirates/0287/2020\|EPI_ISL_698 | ···················· | ···················· | ····················· | ···················· | ···················· | ···················· |
| W | 6427 | hCoV-19/United Arab Emirates/0288/2020\|EPI_ISL_698 | ···················· | ···················· | ····················· | ···················· | ···················· | ···················· |
| W | 6428 | hCoV-19/United Arab Emirates/0289/2020\|EPI_ISL_698 | ···················· | ···················· | ····················· | ···················· | ···················· | ···················· |
| W | 6429 | hCoV-19/United Arab Emirates/0291/2020\|EPI_ISL_698 | ···················· | ···················· | ····················· | ···················· | ···················· | ···················· |
| W | 6430 | hCoV-19/United Arab Emirates/0292/2020\|EPI_ISL_698 | ···················· | ···················· | ····················· | ···················· | ···················· | ···················· |
| W | 6431 | hCoV-19/United Arab Emirates/0293/2020\|EPI_ISL_698 | ···················· | ·A·················· | ····················· | ···················· | ···················· | ···················· |
| W | 6432 | hCoV-19/United Arab Emirates/0294/2020\|EPI_ISL_698 | ···················· | ···················· | ····················· | ···················· | ···················· | ···················· |
| W | 6433 | hCoV-19/United Arab Emirates/0295/2020\|EPI_ISL_698 | ···················· | ···················· | ····················· | ···················· | ···················· | ···················· |

| | | | | | | | |
|---|---|---|---|---|---|---|---|
| W | 6434 hCoV-19/United Arab Emirates/0296/2020\|EPI_ISL_698 | ···················· | ···················· | ···················· | ···················· | ···················· | ···················· |
| W | 6435 hCoV-19/United Arab Emirates/0297/2020\|EPI_ISL_698 | ···················· | ···················· | ···················· | ···················· | ···················· | ···················· |
| W | 6436 hCoV-19/United Arab Emirates/0298/2020\|EPI_ISL_698 | ···················· | ···················· | ···················· | ···················· | ···················· | ···················· |
| W | 6437 hCoV-19/United Arab Emirates/0299/2020\|EPI_ISL_698 | ···················· | ···················· | ···················· | ···················· | ···················· | ···················· |
| W | 6438 hCoV-19/United Arab Emirates/0300/2020\|EPI_ISL_698 | ···················· | ···················· | ···················· | ···················· | ···················· | ···················· |
| W | 6439 hCoV-19/United Arab Emirates/0301/2020\|EPI_ISL_698 | ···················· | ···················· | ···················· | ···················· | ···················· | ···················· |
| W | 6440 hCoV-19/United Arab Emirates/0302/2020\|EPI_ISL_698 | ···················· | ···················· | ···················· | ···················· | ···················· | ···················· |
| W | 6441 hCoV-19/United Arab Emirates/0303/2020\|EPI_ISL_698 | ···················· | ···················· | ···················· | ···················· | ···················· | ···················· |
| W | 6442 hCoV-19/United Arab Emirates/0304/2020\|EPI_ISL_698 | ···················· | ···················· | ···················· | ···················· | ···················· | ···················· |
| W | 6443 hCoV-19/United Arab Emirates/0306/2020\|EPI_ISL_698 | ···················· | ···················· | ···················· | ···················· | ···················· | ···················· |
| W | 6444 hCoV-19/United Arab Emirates/0307/2020\|EPI_ISL_698 | ···················· | ···················· | ···················· | ···················· | ···················· | ···················· |
| W | 6445 hCoV-19/United Arab Emirates/0308/2020\|EPI_ISL_698 | ···················· | ···················· | ···················· | ···················· | ···················· | ···················· |
| W | 6446 hCoV-19/United Arab Emirates/0310/2020\|EPI_ISL_698 | ···················· | ···················· | ···················· | ···················· | ···················· | ···················· |
| W | 6447 hCoV-19/United Arab Emirates/0311/2020\|EPI_ISL_698 | ···················· | ···················· | ···················· | ···················· | ···················· | ···················· |
| W | 6448 hCoV-19/United Arab Emirates/0312/2020\|EPI_ISL_698 | ···················· | ···················· | ···················· | ···················· | ···················· | ···················· |
| W | 6449 hCoV-19/United Arab Emirates/0313/2020\|EPI_ISL_698 | ···················· | ···················· | ···················· | ···················· | ···················· | ···················· |
| W | 6450 hCoV-19/United Arab Emirates/0314/2020\|EPI_ISL_698 | ···················· | ···················· | ···················· | ···················· | ···················· | ···················· |
| W | 6451 hCoV-19/United Arab Emirates/0315/2020\|EPI_ISL_698 | ···················· | ···················· | ···················· | ···················· | ···················· | ···················· |
| W | 6452 hCoV-19/United Arab Emirates/0316/2020\|EPI_ISL_698 | ···················· | ···················· | ···················· | ···················· | ···················· | ···················· |
| W | 6453 hCoV-19/United Arab Emirates/0317/2020\|EPI_ISL_698 | ···················· | ···················· | ···················· | ···················· | ···················· | ···················· |
| W | 6454 hCoV-19/United Arab Emirates/0318/2020\|EPI_ISL_698 | ···················· | ···················· | ···················· | ···················· | ···················· | ···················· |
| W | 6455 hCoV-19/United Arab Emirates/0319/2020\|EPI_ISL_698 | ···················· | ···················· | ···················· | ···················· | ···················· | ···················· |
| W | 6456 hCoV-19/United Arab Emirates/0320/2020\|EPI_ISL_698 | ···················· | ···················· | ···················· | ···················· | ···················· | ···················· |
| W | 6457 hCoV-19/United Arab Emirates/0321/2020\|EPI_ISL_698 | ···················· | ···················· | ···················· | ···················· | ···················· | ···················· |
| W | 6458 hCoV-19/United Arab Emirates/0323/2020\|EPI_ISL_698 | ···················· | ···················· | ···················· | ···················· | ···················· | ···················· |
| W | 6459 hCoV-19/India/MH-ACTREC-114/2020\|EPI_ISL_699770\|20 | ···················· | ···················· | ···················· | ···················· | ···················· | ···················· |
| W | 6461 hCoV-19/India/MH-ACTREC-116/2020\|EPI_ISL_699772\|20 | ···················· | ···················· | ···················· | ···················· | ···················· | ···················· |
| W | 6462 hCoV-19/India/MH-ACTREC-117/2020\|EPI_ISL_699773\|20 | ···················· | ···················· | ···················· | ···················· | ···················· | ···················· |
| W | 6463 hCoV-19/India/MH-ACTREC-118/2020\|EPI_ISL_699774\|20 | ···················· | ···················· | ···················· | ···················· | ···················· | ···················· |
| W | 6464 hCoV-19/India/MH-ACTREC-119/2020\|EPI_ISL_699775\|20 | ···················· | ···················· | ···················· | ···················· | ···················· | ···················· |
| W | 6465 hCoV-19/India/MH-ACTREC-122/2020\|EPI_ISL_699778\|20 | ···················· | ···················· | ···················· | ···················· | ···················· | ···················· |
| W | 6467 hCoV-19/India/MH-ACTREC-124/2020\|EPI_ISL_699780\|20 | ···················· | ···················· | ···················· | ···················· | ···················· | ···················· |
| W | 6468 hCoV-19/India/MH-ACTREC-125/2020\|EPI_ISL_699781\|20 | ···················· | ···················· | ···················· | ···················· | ···················· | ···················· |
| W | 6470 hCoV-19/India/MH-ACTREC-127/2020\|EPI_ISL_699783\|20 | ···················· | ···················· | ···················· | ···················· | ···················· | ···················· |
| W | 6471 hCoV-19/India/MH-ACTREC-128/2020\|EPI_ISL_699784\|20 | ···················· | ···················· | ···················· | ···················· | ···················· | ···················· |
| W | 6472 hCoV-19/India/MH-ACTREC-129/2020\|EPI_ISL_699785\|20 | ···················· | ···················· | ···················· | ···················· | ···················· | ···················· |
| W | 6473 hCoV-19/India/MH-ACTREC-130/2020\|EPI_ISL_699786\|20 | ···················· | ···················· | ···················· | ···················· | ···················· | ···················· |
| W | 6474 hCoV-19/India/MH-ACTREC-131/2020\|EPI_ISL_699787\|20 | ···················· | ···················· | ···················· | ···················· | ···················· | ···················· |
| W | 6475 hCoV-19/India/MH-ACTREC-132/2020\|EPI_ISL_699788\|20 | ···················· | ···················· | ···················· | ···················· | ···················· | ···················· |
| W | 6476 hCoV-19/India/MH-ACTREC-133/2020\|EPI_ISL_699789\|20 | ···················· | ···················· | ···················· | ···················· | ···················· | ···················· |
| W | 6477 hCoV-19/India/MH-ACTREC-134/2020\|EPI_ISL_699790\|20 | ···················· | ···················· | ···················· | ···················· | ···················· | ···················· |
| W | 6478 hCoV-19/India/MH-ACTREC-135/2020\|EPI_ISL_699791\|20 | ···················· | ···················· | ···················· | ···················· | ···················· | ···················· |
| W | 6479 hCoV-19/India/MH-ACTREC-136/2020\|EPI_ISL_699792\|20 | ···················· | ···················· | ···················· | ···················· | ···················· | ···················· |
| W | 6480 hCoV-19/India/MH-ACTREC-137/2020\|EPI_ISL_699793\|20 | ···················· | ···················· | ···················· | ···················· | ···················· | ···················· |
| W | 6481 hCoV-19/India/MH-ACTREC-138/2020\|EPI_ISL_699794\|20 | ···················· | ···················· | ···················· | ···················· | ···················· | ···················· |
| W | 6482 hCoV-19/India/MH-ACTREC-139/2020\|EPI_ISL_699795\|20 | ···················· | ···················· | ···················· | ···················· | ···················· | ···················· |
| W | 6483 hCoV-19/India/MH-ACTREC-140/2020\|EPI_ISL_699796\|20 | ···················· | ···················· | ···················· | ···················· | ···················· | ···················· |
| W | 6484 hCoV-19/India/MH-ACTREC-141/2020\|EPI_ISL_699797\|20 | ···················· | ···················· | ···················· | ···················· | ···················· | ···················· |
| W | 6486 hCoV-19/India/MH-ACTREC-143/2020\|EPI_ISL_699799\|20 | ···················· | ···················· | ···················· | ···················· | ···················· | ···················· |
| W | 6487 hCoV-19/England/ALDP-BB596E/2020\|EPI_ISL_703000\|20 | ···················· | ···················· | ···················· | ···················· | ···················· | ···················· |
| W | 6488 hCoV-19/England/ALDP-BB32A1/2020\|EPI_ISL_703001\|20 | ···················· | ···················· | ···················· | ···················· | ···················· | ···················· |
| W | 6489 hCoV-19/England/ALDP-BB9A58/2020\|EPI_ISL_703003\|20 | ···················· | ·····A·············· | ···················· | ···················· | ···················· | ···················· |
| W | 6490 hCoV-19/England/ALDP-BB34C9/2020\|EPI_ISL_703004\|20 | ···················· | ···················· | ···················· | ···················· | ···················· | ···················· |
| W | 6491 hCoV-19/England/ALDP-BB56D0/2020\|EPI_ISL_703006\|20 | ···················· | ···················· | ···················· | ···················· | ···················· | ···················· |
| W | 6492 hCoV-19/England/ALDP-BB2AF5/2020\|EPI_ISL_703009\|20 | ···················· | ···················· | ···················· | ···················· | ···················· | ···················· |
| W | 6493 hCoV-19/England/ALDP-BB5904/2020\|EPI_ISL_703011\|20 | ···················· | ···················· | ···················· | ···················· | ···················· | ···················· |
| W | 6494 hCoV-19/England/ALDP-BB33F9/2020\|EPI_ISL_703012\|20 | ···················· | ···················· | ···················· | ···················· | ···················· | ···················· |
| W | 6495 hCoV-19/England/ALDP-BB94B4/2020\|EPI_ISL_703016\|20 | ···················· | ···················· | ···················· | ···················· | ···················· | ···················· |
| W | 6496 hCoV-19/England/ALDP-BB2855/2020\|EPI_ISL_703017\|20 | ···················· | ···················· | ···················· | ···················· | ···················· | ··A················· |
| W | 6497 hCoV-19/England/ALDP-BB5DAE/2020\|EPI_ISL_703019\|20 | ···················· | ···················· | ···················· | ···················· | ···················· | ···················· |
| W | 6498 hCoV-19/England/QEUH-BCD329/2020\|EPI_ISL_704300\|20 | ···················· | ·A·················· | ···················· | ···················· | ···················· | ···················· |
| W | 6499 hCoV-19/England/QEUH-BCC16C/2020\|EPI_ISL_704303\|20 | ···················· | ···················· | ···················· | ···················· | ···················· | ···················· |
| W | 6500 hCoV-19/England/QEUH-BCDDFE/2020\|EPI_ISL_704304\|20 | ···················· | ···················· | ···················· | ···················· | ···················· | ···················· |
| W | 6501 hCoV-19/England/QEUH-BCCECF/2020\|EPI_ISL_704306\|20 | ···················· | ···················· | ···················· | ···················· | ···················· | ···················· |
| W | 6502 hCoV-19/England/QEUH-BCCF80/2020\|EPI_ISL_704310\|20 | ···················· | ···················· | ···················· | ···················· | ···················· | ···················· |
| W | 6503 hCoV-19/England/QEUH-BCCDC2/2020\|EPI_ISL_704313\|20 | ···················· | ···················· | ···················· | ···················· | ···················· | ···················· |
| W | 6504 hCoV-19/England/QEUH-BCD383/2020\|EPI_ISL_704314\|20 | ···················· | ···················· | ···················· | ···················· | ···················· | ···················· |
| W | 6505 hCoV-19/England/QEUH-BCCB04/2020\|EPI_ISL_704316\|20 | ···················· | ···················· | ···················· | ···················· | ···················· | ···················· |
| W | 6506 hCoV-19/England/QEUH-BCC21E/2020\|EPI_ISL_704318\|20 | ···················· | ···················· | ···················· | ···················· | ···················· | ···················· |
| W | 6507 hCoV-19/England/QEUH-BCE883/2020\|EPI_ISL_704330\|20 | ···················· | ···················· | ···················· | ···················· | ···················· | ···················· |
| W | 6508 hCoV-19/England/QEUH-BCE373/2020\|EPI_ISL_704335\|20 | ···················· | ···················· | ···················· | ···················· | ···················· | ···················· |
| W | 6509 hCoV-19/England/QEUH-BCE319/2020\|EPI_ISL_704337\|20 | ···················· | ···················· | ···················· | ···················· | ···················· | ···················· |
| W | 6510 hCoV-19/England/QEUH-BCD462/2020\|EPI_ISL_704338\|20 | ···················· | ···················· | ···················· | ···················· | ···················· | ···················· |
| W | 6511 hCoV-19/England/QEUH-BCD9CD/2020\|EPI_ISL_704340\|20 | ···················· | ···················· | ···················· | ···················· | ···················· | ···················· |
| W | 6512 hCoV-19/England/QEUH-BCB5BC/2020\|EPI_ISL_704342\|20 | ···················· | ···················· | ···················· | ···················· | ·····T·············· | ···················· |
| W | 6513 hCoV-19/England/QEUH-BCBC11/2020\|EPI_ISL_704343\|20 | ···················· | ···················· | ···················· | ···················· | ···················· | ···················· |
| W | 6514 hCoV-19/England/QEUH-BCE2D0/2020\|EPI_ISL_704345\|20 | ···················· | ···················· | ···················· | ···················· | ···················· | ···················· |
| W | 6515 hCoV-19/England/QEUH-BCD74B/2020\|EPI_ISL_704346\|20 | ···················· | ···················· | ···················· | ···················· | ·····T·············· | ···················· |
| W | 6516 hCoV-19/England/QEUH-BCD70F/2020\|EPI_ISL_704348\|20 | ···················· | ···················· | ···················· | ···················· | ···················· | ···················· |
| W | 6517 hCoV-19/England/QEUH-BCD6A8/2020\|EPI_ISL_704349\|20 | ···················· | ···················· | ···················· | ···················· | ···················· | ···················· |

| | | | | | | | | |
|---|---|---|---|---|---|---|---|---|
| W | 6518 | hCoV-19/England/CAMC-BBC83D/2020\|EPI_ISL_705600\|20 | ···················· | ···················· | ····················· | ···················· | ···················· | ···················· |
| W | 6519 | hCoV-19/England/CAMC-BBC84C/2020\|EPI_ISL_705601\|20 | ···················· | ···················· | ····················· | ···················· | ···················· | ···················· |
| W | 6520 | hCoV-19/England/CAMC-BBCA46/2020\|EPI_ISL_705602\|20 | ···················· | ···················· | ····················· | ···················· | ···················· | ···················· |
| W | 6521 | hCoV-19/England/CAMC-BBCA55/2020\|EPI_ISL_705603\|20 | ···················· | ···················· | ····················· | ···················· | ···················· | ···················· |
| W | 6522 | hCoV-19/England/CAMC-BBCA91/2020\|EPI_ISL_705604\|20 | ···················· | ···················· | ····················· | ···················· | ···················· | ···················· |
| W | 6523 | hCoV-19/England/CAMC-BBCABF/2020\|EPI_ISL_705605\|20 | ···················· | ···················· | ····················· | ···················· | ···················· | ···················· |
| W | 6524 | hCoV-19/England/CAMC-BBCADD/2020\|EPI_ISL_705606\|20 | ···················· | ···················· | ····················· | ···················· | ······T············· | ···················· |
| W | 6525 | hCoV-19/England/CAMC-BBCAEC/2020\|EPI_ISL_705607\|20 | ········T··········· | ···················· | ····················· | ···················· | ···················· | ···················· |
| W | 6526 | hCoV-19/England/CAMC-BBCAFB/2020\|EPI_ISL_705608\|20 | ···················· | ···················· | ····················· | ···················· | ···················· | ···················· |
| W | 6527 | hCoV-19/England/CAMC-BBCB07/2020\|EPI_ISL_705609\|20 | ···················· | ···················· | ····················· | ···················· | ···················· | ···················· |
| W | 6528 | hCoV-19/England/CAMC-BBCB25/2020\|EPI_ISL_705610\|20 | ···················· | ···················· | ····················· | ···················· | ···················· | ···················· |
| W | 6529 | hCoV-19/England/CAMC-BBCB34/2020\|EPI_ISL_705611\|20 | ········T··········· | ···················· | ····················· | ···················· | ···················· | ···················· |
| W | 6530 | hCoV-19/England/CAMC-BBCB43/2020\|EPI_ISL_705612\|20 | ···················· | ···················· | ····················· | ···················· | ···················· | ···················· |
| W | 6531 | hCoV-19/England/CAMC-BBCE59/2020\|EPI_ISL_705613\|20 | ···················· | ···················· | ····················· | ···················· | ···················· | ···················· |
| W | 6532 | hCoV-19/England/CAMC-BBCF83/2020\|EPI_ISL_705614\|20 | ···················· | ···················· | ····················· | ···················· | ···················· | ···················· |
| W | 6533 | hCoV-19/England/CAMC-BBDB15/2020\|EPI_ISL_705615\|20 | ···················· | ···················· | ····················· | ···················· | ···················· | ···················· |
| W | 6534 | hCoV-19/England/CAMC-BBDB33/2020\|EPI_ISL_705616\|20 | ···················· | ···················· | ····················· | ···················· | ···················· | ···················· |
| W | 6535 | hCoV-19/England/CAMC-BBDB51/2020\|EPI_ISL_705617\|20 | ···················· | ···················· | ····················· | ···················· | ···················· | ···················· |
| W | 6536 | hCoV-19/England/CAMC-BBDB7F/2020\|EPI_ISL_705618\|20 | ···················· | ···················· | ····················· | ···················· | ···················· | ···················· |
| W | 6537 | hCoV-19/England/CAMC-BBDD00/2020\|EPI_ISL_705620\|20 | ···················· | ···················· | ····················· | ···················· | ···················· | ···················· |
| W | 6538 | hCoV-19/England/CAMC-BBDD3D/2020\|EPI_ISL_705621\|20 | ···················· | ···················· | ····················· | ···················· | ···················· | ···················· |
| W | 6539 | hCoV-19/England/CAMC-BBDDA6/2020\|EPI_ISL_705622\|20 | ···················· | ···················· | ····················· | ···················· | ···················· | ···················· |
| W | 6540 | hCoV-19/England/CAMC-BBDDD3/2020\|EPI_ISL_705623\|20 | ···················· | ···················· | ····················· | ···················· | ···················· | ···················· |
| W | 6541 | hCoV-19/England/CAMC-BBDDF1/2020\|EPI_ISL_705624\|20 | ···················· | ···················· | ····················· | ···················· | ······T············· | ···················· |
| W | 6542 | hCoV-19/England/CAMC-BBDE2B/2020\|EPI_ISL_705625\|20 | ···················· | ···················· | ····················· | ···················· | ···················· | ···················· |
| W | 6543 | hCoV-19/England/CAMC-BBDE49/2020\|EPI_ISL_705626\|20 | ···················· | ···················· | ····················· | ···················· | ······T············· | ···················· |
| W | 6544 | hCoV-19/England/CAMC-BBDE58/2020\|EPI_ISL_705627\|20 | ···················· | ···················· | ····················· | ···················· | ······T············· | ···················· |
| W | 6545 | hCoV-19/England/CAMC-BBE455/2020\|EPI_ISL_705628\|20 | ···················· | ···················· | ····················· | ···················· | ···················· | ···················· |
| W | 6546 | hCoV-19/England/CAMC-BC17CF/2020\|EPI_ISL_705629\|20 | ···················· | ···················· | ····················· | ···················· | ···················· | ···················· |
| W | 6547 | hCoV-19/England/CAMC-BC1835/2020\|EPI_ISL_705630\|20 | ···················· | ···················· | ····················· | ···················· | ···················· | ···················· |
| W | 6548 | hCoV-19/England/CAMC-BC1B4B/2020\|EPI_ISL_705631\|20 | ···················· | ···················· | ····················· | ···················· | ···················· | ···················· |
| W | 6549 | hCoV-19/England/CAMC-BC1DBE/2020\|EPI_ISL_705632\|20 | ···················· | ···················· | ····················· | ···················· | ···················· | ···················· |
| W | 6550 | hCoV-19/England/CAMC-BC1E9D/2020\|EPI_ISL_705633\|20 | ···················· | ···················· | ····················· | ···················· | ···················· | ···················· |
| W | 6551 | hCoV-19/England/CAMC-BC1EBB/2020\|EPI_ISL_705634\|20 | ···················· | ···················· | ····················· | ···················· | ···················· | ···················· |
| W | 6552 | hCoV-19/England/CAMC-BC35B4/2020\|EPI_ISL_705635\|20 | ···················· | ···················· | ····················· | ···················· | ···················· | ···················· |
| W | 6553 | hCoV-19/Scotland/CVR5761/2020\|EPI_ISL_705636\|2020- | ···················· | ···················· | ····················· | ···················· | ···················· | ···················· |
| W | 6554 | hCoV-19/Scotland/CVR5762/2020\|EPI_ISL_705637\|2020- | ···················· | ···················· | ····················· | ···················· | ···················· | ···················· |
| W | 6555 | hCoV-19/Scotland/CVR5768/2020\|EPI_ISL_705638\|2020- | ···················· | ···················· | ····················· | ···················· | ···················· | ···················· |
| W | 6556 | hCoV-19/Scotland/CVR5769/2020\|EPI_ISL_705639\|2020- | ···················· | ···················· | ····················· | ···················· | ···················· | ···················· |
| W | 6557 | hCoV-19/Scotland/CVR5770/2020\|EPI_ISL_705640\|2020- | ···················· | ···················· | ····················· | ···················· | ···················· | ···················· |
| W | 6558 | hCoV-19/Scotland/CVR5879/2020\|EPI_ISL_705641\|2020- | ···················· | ···················· | ····················· | ···················· | ···················· | ···················· |
| W | 6559 | hCoV-19/Scotland/CVR5880/2020\|EPI_ISL_705642\|2020- | ···················· | ···················· | ····················· | ···················· | ···················· | ···················· |
| W | 6560 | hCoV-19/Scotland/CVR5881/2020\|EPI_ISL_705643\|2020- | ···················· | ···················· | ····················· | ···················· | ···················· | ···················· |
| W | 6561 | hCoV-19/Scotland/CVR5882/2020\|EPI_ISL_705644\|2020- | ···················· | ···················· | ····················· | ···················· | ···················· | ···················· |
| W | 6562 | hCoV-19/Scotland/CVR5883/2020\|EPI_ISL_705645\|2020- | ···················· | ···················· | ····················· | ···················· | ···················· | ···················· |
| W | 6563 | hCoV-19/Scotland/CVR5884/2020\|EPI_ISL_705646\|2020- | ···················· | ···················· | ····················· | ···················· | ···················· | ···················· |
| W | 6564 | hCoV-19/Scotland/CVR5887/2020\|EPI_ISL_705647\|2020- | ···················· | ···················· | ····················· | ···················· | ···················· | ···················· |
| W | 6565 | hCoV-19/Scotland/CVR5902/2020\|EPI_ISL_705660\|2020- | ···················· | ···················· | ····················· | ···················· | ···················· | ···················· |
| W | 6566 | hCoV-19/Scotland/CVR5903/2020\|EPI_ISL_705661\|2020- | ···················· | ···················· | ····················· | ···················· | ···················· | ···················· |
| W | 6567 | hCoV-19/Scotland/CVR5904/2020\|EPI_ISL_705662\|2020- | ···················· | ···················· | ····················· | ···················· | ···················· | ···················· |
| W | 6568 | hCoV-19/Scotland/CVR5905/2020\|EPI_ISL_705663\|2020- | ···················· | ···················· | ····················· | ···················· | ···················· | ···················· |
| W | 6569 | hCoV-19/Scotland/CVR5906/2020\|EPI_ISL_705664\|2020- | ···················· | ···················· | ····················· | ···················· | ···················· | ···················· |
| W | 6570 | hCoV-19/Scotland/CVR5909/2020\|EPI_ISL_705665\|2020- | ···················· | ···················· | ····················· | ···················· | ···················· | ···················· |
| W | 6571 | hCoV-19/Scotland/CVR5910/2020\|EPI_ISL_705666\|2020- | ···················· | ···················· | ····················· | ···················· | ···················· | ···················· |
| W | 6572 | hCoV-19/Scotland/CVR5914/2020\|EPI_ISL_705667\|2020- | ···················· | ···················· | ····················· | ···················· | ···················· | ···················· |
| W | 6573 | hCoV-19/Scotland/CVR5915/2020\|EPI_ISL_705668\|2020- | ···················· | ···················· | ····················· | ···················· | ···················· | ···················· |
| W | 6574 | hCoV-19/Scotland/CVR5918/2020\|EPI_ISL_705669\|2020- | ···················· | ···················· | ····················· | ···················· | ···················· | ···················· |
| W | 6575 | hCoV-19/Scotland/CVR5920/2020\|EPI_ISL_705670\|2020- | ···················· | ···················· | ····················· | ···················· | ···················· | ···················· |
| W | 6576 | hCoV-19/Scotland/CVR5922/2020\|EPI_ISL_705671\|2020- | ···················· | ···················· | ····················· | ···················· | ···················· | ···················· |
| W | 6577 | hCoV-19/Scotland/CVR5923/2020\|EPI_ISL_705672\|2020- | ···················· | ···················· | ····················· | ···················· | ···················· | ···················· |
| W | 6578 | hCoV-19/Scotland/CVR5924/2020\|EPI_ISL_705673\|2020- | ···················· | ···················· | ····················· | ···················· | ···················· | ···················· |
| W | 6579 | hCoV-19/Scotland/CVR5925/2020\|EPI_ISL_705674\|2020- | ···················· | ···················· | ····················· | ···················· | ···················· | ···················· |
| W | 6580 | hCoV-19/Scotland/CVR5926/2020\|EPI_ISL_705675\|2020- | ···················· | ···················· | ····················· | ···················· | ···················· | ···················· |
| W | 6581 | hCoV-19/Scotland/CVR5930/2020\|EPI_ISL_705676\|2020- | ···················· | ···················· | ····················· | ···················· | ···················· | ···················· |
| W | 6582 | hCoV-19/Scotland/CVR5932/2020\|EPI_ISL_705677\|2020- | ···················· | ···················· | ····················· | ···················· | ···················· | ···················· |
| W | 6583 | hCoV-19/Scotland/CVR5933/2020\|EPI_ISL_705678\|2020- | ···················· | ···················· | ····················· | ···················· | ···················· | ···················· |
| W | 6584 | hCoV-19/Scotland/CVR5934/2020\|EPI_ISL_705679\|2020- | ···················· | ···················· | ····················· | ···················· | ···················· | ···················· |
| W | 6585 | hCoV-19/England/QEUH-A370F7/2020\|EPI_ISL_706928\|20 | ···················· | ···················· | ····················· | ···················· | ···················· | ···················· |
| W | 6586 | hCoV-19/England/QEUH-A37103/2020\|EPI_ISL_706929\|20 | ···················· | ···················· | ····················· | ···················· | ···················· | ···················· |
| W | 6587 | hCoV-19/England/QEUH-A619CC/2020\|EPI_ISL_706930\|20 | ···················· | ·A·················· | ····················· | ···················· | ···················· | ···················· |
| W | 6588 | hCoV-19/England/QEUH-A619DB/2020\|EPI_ISL_706931\|20 | ···················· | ···················· | ····················· | ···················· | ···················· | ···················· |
| W | 6589 | hCoV-19/England/QEUH-A619EA/2020\|EPI_ISL_706932\|20 | ···················· | ···················· | ····················· | ···················· | ···················· | ···················· |
| W | 6590 | hCoV-19/England/QEUH-A619F9/2020\|EPI_ISL_706933\|20 | ···················· | ···················· | ····················· | ···················· | ···················· | ···················· |
| W | 6591 | hCoV-19/England/QEUH-A61A05/2020\|EPI_ISL_706934\|20 | ···················· | ···················· | ····················· | ···················· | ···················· | ···················· |
| W | 6592 | hCoV-19/England/QEUH-A61A32/2020\|EPI_ISL_706935\|20 | ···················· | ···················· | ····················· | ···················· | ······T············· | ···················· |
| W | 6593 | hCoV-19/England/QEUH-A61A50/2020\|EPI_ISL_706936\|20 | ···················· | ···················· | ····················· | ···················· | ···················· | ···················· |
| W | 6594 | hCoV-19/England/QEUH-A61A6F/2020\|EPI_ISL_706937\|20 | ···················· | ···················· | ····················· | ···················· | ···················· | ···················· |
| W | 6595 | hCoV-19/England/QEUH-A61E18/2020\|EPI_ISL_706938\|20 | ···················· | ···················· | ····················· | ···················· | ···················· | ···················· |
| W | 6596 | hCoV-19/England/QEUH-A61E36/2020\|EPI_ISL_706939\|20 | ···················· | ···················· | ····················· | ···················· | ···················· | ···················· |
| W | 6597 | hCoV-19/England/QEUH-A61ECD/2020\|EPI_ISL_706940\|20 | ···················· | ···················· | ····················· | ···················· | ···················· | ···················· |

| | | | | | | | | |
|---|---|---|---|---|---|---|---|---|
| W | 6598 hCoV-19/England/QEUH-A61EDC/2020\|EPI_ISL_706941\|20 | ···················· | ···················· | ···················· | ···················· | ···················· | ···················· | |
| W | 6599 hCoV-19/England/QEUH-A61EEB/2020\|EPI_ISL_706942\|20 | ···················· | ···················· | ···················· | ···················· | ···················· | ···················· | |
| W | 6600 hCoV-19/England/QEUH-A61F15/2020\|EPI_ISL_706943\|20 | ···················· | ···················· | ···················· | ···················· | ···················· | ···················· | |
| W | 6601 hCoV-19/England/QEUH-A61F24/2020\|EPI_ISL_706944\|20 | ···················· | ···················· | ···················· | ···················· | ···················· | ···················· | |
| W | 6602 hCoV-19/England/QEUH-A61F33/2020\|EPI_ISL_706945\|20 | ···················· | ···················· | ···················· | ···················· | ···················· | ···················· | |
| W | 6603 hCoV-19/England/QEUH-A61F42/2020\|EPI_ISL_706946\|20 | ···················· | ···················· | ···················· | ···················· | ···················· | ···················· | |
| W | 6604 hCoV-19/England/QEUH-A61F51/2020\|EPI_ISL_706947\|20 | ···················· | ···················· | ···················· | ···················· | ···················· | ···················· | |
| W | 6605 hCoV-19/England/QEUH-A61F60/2020\|EPI_ISL_706948\|20 | ···················· | ···················· | ···················· | ···················· | ···················· | ···················· | |
| W | 6606 hCoV-19/England/QEUH-A61F7F/2020\|EPI_ISL_706949\|20 | ···················· | ···················· | ···················· | ···················· | ···················· | ···················· | |
| W | 6607 hCoV-19/England/QEUH-A61F8E/2020\|EPI_ISL_706950\|20 | ···················· | ···················· | ···················· | ···················· | ···················· | ···················· | |
| W | 6608 hCoV-19/England/QEUH-A61FCA/2020\|EPI_ISL_706951\|20 | ···················· | ···················· | ···················· | ···················· | ···················· | ···················· | |
| W | 6609 hCoV-19/England/QEUH-A61FF7/2020\|EPI_ISL_706952\|20 | ···················· | ···················· | ···················· | ···················· | ···················· | ···················· | |
| W | 6610 hCoV-19/England/QEUH-A62002/2020\|EPI_ISL_706953\|20 | ···················· | ···················· | ···················· | ···················· | ···················· | ···················· | |
| W | 6611 hCoV-19/England/QEUH-A6207B/2020\|EPI_ISL_706954\|20 | ···················· | ···················· | ···················· | ···················· | ···················· | ···················· | |
| W | 6612 hCoV-19/England/QEUH-A62099/2020\|EPI_ISL_706955\|20 | ···················· | ·A·················· | ···················· | ···················· | ···················· | ···················· | |
| W | 6613 hCoV-19/England/QEUH-A620A8/2020\|EPI_ISL_706956\|20 | ···················· | ···················· | ···················· | ···················· | ···················· | ···················· | |
| W | 6614 hCoV-19/England/QEUH-A620C6/2020\|EPI_ISL_706957\|20 | ···················· | ·A·················· | ···················· | ···················· | ···················· | ··A················· | |
| W | 6615 hCoV-19/England/QEUH-A620E4/2020\|EPI_ISL_706958\|20 | ···················· | ·A·················· | ···················· | ···················· | ···················· | ··A················· | |
| W | 6616 hCoV-19/England/QEUH-A6210F/2020\|EPI_ISL_706959\|20 | ···················· | ···················· | ···················· | ···················· | ···················· | ···················· | |
| W | 6617 hCoV-19/England/QEUH-A6214B/2020\|EPI_ISL_706960\|20 | ···················· | ···················· | ···················· | ···················· | ···················· | ···················· | |
| W | 6618 hCoV-19/England/QEUH-A621A5/2020\|EPI_ISL_706961\|20 | ···················· | ···················· | ···················· | ···················· | ···················· | ···················· | |
| W | 6619 hCoV-19/England/QEUH-A621B4/2020\|EPI_ISL_706962\|20 | ···················· | ···················· | ···················· | ···················· | ···················· | ···················· | |
| W | 6620 hCoV-19/Scotland/QEUH-A621C3/2020\|EPI_ISL_706963\|2 | ···················· | ···················· | ···················· | ···················· | ·····T·············· | ···················· | |
| W | 6621 hCoV-19/England/QEUH-A622C0/2020\|EPI_ISL_706964\|20 | ···················· | ···················· | ···················· | ···················· | ···················· | ···················· | |
| W | 6622 hCoV-19/Scotland/QEUH-A62406/2020\|EPI_ISL_706965\|2 | ···················· | ···················· | ···················· | ···················· | ·····T·············· | ···················· | |
| W | 6623 hCoV-19/England/QEUH-A624F7/2020\|EPI_ISL_706966\|20 | ···················· | ···················· | ···················· | ···················· | ···················· | ···················· | |
| W | 6624 hCoV-19/England/QEUH-A62512/2020\|EPI_ISL_706967\|20 | ···················· | ···················· | ···················· | ···················· | ···················· | ···················· | |
| W | 6625 hCoV-19/Scotland/QEUH-A62530/2020\|EPI_ISL_706968\|2 | ···················· | ···················· | ···················· | ···················· | ···················· | ···················· | |
| W | 6626 hCoV-19/England/QEUH-A6254F/2020\|EPI_ISL_706969\|20 | ···················· | ···················· | ···················· | ···················· | ···················· | ···················· | |
| W | 6627 hCoV-19/England/QEUH-A6257C/2020\|EPI_ISL_706970\|20 | ···················· | ·A·················· | ···················· | ···················· | ···················· | ··A················· | |
| W | 6628 hCoV-19/Wales/QEUH-A625A9/2020\|EPI_ISL_706971\|2020 | ···················· | ···················· | ···················· | ···················· | ···················· | ···················· | |
| W | 6629 hCoV-19/England/QEUH-A625C7/2020\|EPI_ISL_706972\|20 | ···················· | ···················· | ···················· | ···················· | ···················· | ···················· | |
| W | 6630 hCoV-19/England/QEUH-A62600/2020\|EPI_ISL_706973\|20 | ···················· | ···················· | ···················· | ···················· | ···················· | ···················· | |
| W | 6631 hCoV-19/England/QEUH-A8A70F/2020\|EPI_ISL_706974\|20 | ···················· | ···················· | ···················· | ···················· | ···················· | ···················· | |
| W | 6632 hCoV-19/England/SHEF-10C912B/2020\|EPI_ISL_706975\|2 | ···················· | ···················· | ···················· | ···················· | ···················· | ···················· | |
| W | 6633 hCoV-19/England/SHEF-10C9C93/2020\|EPI_ISL_706976\|2 | ···················· | ···················· | ···················· | ···················· | ···················· | ···················· | |
| W | 6634 hCoV-19/England/SHEF-10CAC65/2020\|EPI_ISL_706977\|2 | ···················· | ···················· | ···················· | ···················· | ··A················· | ···················· | |
| W | 6635 hCoV-19/England/SHEF-10CDDBA/2020\|EPI_ISL_706978\|2 | ···················· | ···················· | ···················· | ···················· | ···················· | ···················· | |
| W | 6636 hCoV-19/England/SHEF-10CEDE6/2020\|EPI_ISL_706979\|2 | ···················· | ···················· | ···················· | ···················· | ···················· | ···················· | |
| W | 6637 hCoV-19/England/SHEF-10D85E1/2020\|EPI_ISL_706990\|2 | ···················· | ···················· | ···················· | ···················· | ···················· | ···················· | |
| W | 6638 hCoV-19/England/SHEF-C1083/2020\|EPI_ISL_706991\|202 | ···················· | ···················· | ···················· | ···················· | ···················· | ···················· | |
| W | 6639 hCoV-19/England/SHEF-C19C4/2020\|EPI_ISL_706992\|202 | ···················· | ···················· | ···················· | ···················· | ···················· | ···················· | |
| W | 6640 hCoV-19/England/SHEF-C4466/2020\|EPI_ISL_706993\|202 | ···················· | ···················· | ···················· | ···················· | ···················· | ···················· | |
| W | 6641 hCoV-19/England/SHEF-C48A6/2020\|EPI_ISL_706994\|202 | ···················· | ···················· | ···················· | ···················· | ···················· | ···················· | |
| W | 6642 hCoV-19/England/SHEF-C516E/2020\|EPI_ISL_706995\|202 | ···················· | ···················· | ···················· | ···················· | ···················· | ···················· | |
| W | 6643 hCoV-19/England/SHEF-C617C/2020\|EPI_ISL_706996\|202 | ···················· | ···················· | ···················· | ···················· | ···················· | ···················· | |
| W | 6644 hCoV-19/England/SHEF-C7B22/2020\|EPI_ISL_706997\|202 | ···················· | ···················· | ···················· | ···················· | ···················· | ···················· | |
| W | 6645 hCoV-19/England/SHEF-C880C/2020\|EPI_ISL_706998\|202 | ···················· | ···················· | ···················· | ···················· | ···················· | ···················· | |
| W | 6646 hCoV-19/England/SHEF-CA98F/2020\|EPI_ISL_706999\|202 | ···················· | ···················· | ···················· | ···················· | ···················· | ···················· | |
| W | 6647 hCoV-19/USA/CO-CDPHE-2004220758/2020\|EPI_ISL_71019 | ···················· | ···················· | ···················· | ···················· | ·NNN················ | ···················· | 1 |
| W | 6648 hCoV-19/USA/CO-CDPHE-2004220761/2020\|EPI_ISL_71019 | ···················· | ···················· | ···················· | ···················· | ···················· | ···················· | |
| W | 6650 hCoV-19/USA/CO-CDPHE-2004220764/2020\|EPI_ISL_71019 | ···················· | ···················· | ···················· | ···················· | NNN················· | ···················· | 1 |
| W | 6651 hCoV-19/USA/CO-CDPHE-2004220766/2020\|EPI_ISL_71019 | ···················· | ···················· | ···················· | ···················· | ···················· | ···················· | |
| W | 6652 hCoV-19/USA/CO-CDPHE-2004220771/2020\|EPI_ISL_71019 | ···················· | ···················· | ···················· | ···················· | ···················· | ···················· | |
| W | 6655 hCoV-19/USA/CO-CDPHE-2004220785/2020\|EPI_ISL_71019 | ···················· | ···················· | ···················· | ···················· | ···················· | ···················· | |
| W | 6656 hCoV-19/USA/CO-CDPHE-2004220869/2020\|EPI_ISL_71019 | ···················· | ···················· | ···················· | ···················· | ···················· | ···················· | |
| W | 6657 hCoV-19/USA/AZ-TG405413/2020\|EPI_ISL_694872\|2020-0 | ···················· | ···················· | ···················· | ···················· | ···················· | ···················· | |
| W | 6658 hCoV-19/USA/AZ-TG407779/2020\|EPI_ISL_694881\|2020-0 | ···················· | ···················· | ···················· | ···················· | ···················· | ···················· | |
| W | 6659 hCoV-19/USA/AZ-TG408005/2020\|EPI_ISL_694884\|2020-0 | ···················· | ···················· | ···················· | ···················· | ···················· | ···················· | |
| W | 6660 hCoV-19/USA/AZ-TG408122/2020\|EPI_ISL_694885\|2020-0 | ···················· | ·A·················· | ···················· | ···················· | ···················· | ···················· | |
| W | 6661 hCoV-19/USA/AZ-TG408124/2020\|EPI_ISL_694887\|2020-0 | ···················· | ·A·················· | ···················· | ···················· | ···················· | ···················· | |
| W | 6662 hCoV-19/USA/AZ-TG408125/2020\|EPI_ISL_694888\|2020-0 | ···················· | ·A·················· | ···················· | ···················· | ···················· | ···················· | |
| W | 6663 hCoV-19/USA/AZ-TG408126/2020\|EPI_ISL_694889\|2020-0 | ···················· | ···················· | ···················· | ···················· | ···················· | ···················· | |
| W | 6664 hCoV-19/USA/AZ-TG408127/2020\|EPI_ISL_694890\|2020-0 | ···················· | ···················· | ···················· | ···················· | ···················· | ···················· | |
| W | 6665 hCoV-19/USA/AZ-TG408128/2020\|EPI_ISL_694891\|2020-0 | ···················· | ···················· | ···················· | ···················· | ···················· | ···················· | |
| W | 6666 hCoV-19/USA/AZ-TG408130/2020\|EPI_ISL_694892\|2020-0 | ···················· | ···················· | ···················· | ···················· | ···················· | ···················· | |
| W | 6667 hCoV-19/USA/AZ-TG408133/2020\|EPI_ISL_694893\|2020-0 | ···················· | ···················· | ···················· | ···················· | ···················· | ···················· | |
| W | 6668 hCoV-19/USA/AZ-TG408136/2020\|EPI_ISL_694895\|2020-0 | ···················· | ···················· | ···················· | ···················· | ···················· | ···················· | |
| W | 6669 hCoV-19/USA/AZ-TG408140/2020\|EPI_ISL_694897\|2020-0 | ···················· | ···················· | ···················· | ···················· | ···················· | ···················· | |
| W | 6670 hCoV-19/USA/AZ-TG408142/2020\|EPI_ISL_694898\|2020-0 | ···················· | ···················· | ···················· | ···················· | ···················· | ···················· | |
| W | 6671 hCoV-19/USA/AZ-TG408146/2020\|EPI_ISL_694901\|2020-0 | ···················· | ···················· | ···················· | ···················· | ···················· | ···················· | |
| W | 6672 hCoV-19/USA/AZ-TG408154/2020\|EPI_ISL_694906\|2020-0 | ···················· | ·A·················· | ···················· | ···················· | ···················· | ···················· | |
| W | 6673 hCoV-19/USA/AZ-TG408155/2020\|EPI_ISL_694907\|2020-0 | ···················· | ·A·················· | ···················· | ···················· | ···················· | ···················· | |
| W | 6674 hCoV-19/USA/AZ-TG408156/2020\|EPI_ISL_694908\|2020-0 | ···················· | ·A·················· | ···················· | ···················· | ···················· | ···················· | |
| W | 6675 hCoV-19/USA/AZ-TG408157/2020\|EPI_ISL_694909\|2020-0 | ···················· | ·············A······ | ···················· | ···················· | ···················· | ···················· | |
| W | 6676 hCoV-19/United Arab Emirates/0214/2020\|EPI_ISL_698 | ···················· | ···················· | ···················· | ···················· | ···················· | ···················· | |
| W | 6677 hCoV-19/United Arab Emirates/0215/2020\|EPI_ISL_698 | ···················· | ···················· | ···················· | ···················· | ···················· | ···················· | |
| W | 6678 hCoV-19/United Arab Emirates/0216/2020\|EPI_ISL_698 | ···················· | ···················· | ···················· | ···················· | ···················· | ···················· | |
| W | 6679 hCoV-19/United Arab Emirates/0217/2020\|EPI_ISL_698 | ···················· | ···················· | ···················· | ···················· | ···················· | ···················· | |
| W | 6680 hCoV-19/United Arab Emirates/0218/2020\|EPI_ISL_698 | ···················· | ···················· | ···················· | ···················· | ···················· | ···················· | |

| | | | | | | | |
|---|---|---|---|---|---|---|---|
| W | 6681 hCoV-19/United Arab Emirates/0219/2020\|EPI_ISL_698 | ···················· | ···················· | ···················· | ···················· | ···················· | ···················· |
| W | 6682 hCoV-19/United Arab Emirates/0220/2020\|EPI_ISL_698 | ···················· | ···················· | ···················· | ···················· | ···················· | ···················· |
| W | 6683 hCoV-19/United Arab Emirates/0221/2020\|EPI_ISL_698 | ···················· | ···················· | ···················· | ···················· | ···················· | ···················· |
| W | 6684 hCoV-19/United Arab Emirates/0223/2020\|EPI_ISL_698 | ···················· | ···················· | ···················· | ···················· | ···················· | ···················· |
| W | 6685 hCoV-19/United Arab Emirates/0224/2020\|EPI_ISL_698 | ···················· | ···················· | ···················· | ···················· | ···················· | ···················· |
| W | 6686 hCoV-19/United Arab Emirates/0226/2020\|EPI_ISL_698 | ···················· | ···················· | ···················· | ···················· | ···················· | ···················· |
| W | 6687 hCoV-19/United Arab Emirates/0227/2020\|EPI_ISL_698 | ···················· | ···················· | ···················· | ···················· | ···················· | ···················· |
| W | 6688 hCoV-19/United Arab Emirates/0228/2020\|EPI_ISL_698 | ···················· | ···················· | ···················· | ···················· | ···················· | ···················· |
| W | 6689 hCoV-19/United Arab Emirates/0229/2020\|EPI_ISL_698 | ···················· | ···················· | ···················· | ···················· | ···················· | ···················· |
| W | 6690 hCoV-19/United Arab Emirates/0230/2020\|EPI_ISL_698 | ···················· | ···················· | ···················· | ···················· | ···················· | ···················· |
| W | 6691 hCoV-19/United Arab Emirates/0231/2020\|EPI_ISL_698 | ···················· | ···················· | ···················· | ···················· | ···················· | ···················· |
| W | 6692 hCoV-19/United Arab Emirates/0232/2020\|EPI_ISL_698 | ···················· | ···················· | ···················· | ···················· | ···················· | ···················· |
| W | 6693 hCoV-19/United Arab Emirates/0233/2020\|EPI_ISL_698 | ···················· | ···················· | ···················· | ···················· | ···················· | ···················· |
| W | 6694 hCoV-19/United Arab Emirates/0234/2020\|EPI_ISL_698 | ···················· | ·A·················· | ···················· | ···················· | ···················· | ···················· |
| W | 6695 hCoV-19/United Arab Emirates/0237/2020\|EPI_ISL_698 | ···················· | ···················· | ···················· | ···················· | ···················· | ···················· |
| W | 6696 hCoV-19/United Arab Emirates/0238/2020\|EPI_ISL_698 | ···················· | ···················· | ···················· | ···················· | ···················· | ···················· |
| W | 6697 hCoV-19/United Arab Emirates/0239/2020\|EPI_ISL_698 | ···················· | ···················· | ···················· | ···················· | ···················· | ···················· |
| W | 6698 hCoV-19/United Arab Emirates/0240/2020\|EPI_ISL_698 | ···················· | ···················· | ···················· | ···················· | ···················· | ···················· |
| W | 6699 hCoV-19/United Arab Emirates/0241/2020\|EPI_ISL_698 | ···················· | ···················· | ···················· | ···················· | ···················· | ···················· |
| W | 6700 hCoV-19/United Arab Emirates/0242/2020\|EPI_ISL_698 | ···················· | ···················· | ···················· | ···················· | ···················· | ···················· |
| W | 6701 hCoV-19/United Arab Emirates/0243/2020\|EPI_ISL_698 | ···················· | ···················· | ···················· | ···················· | ···················· | ···················· |
| W | 6702 hCoV-19/United Arab Emirates/0244/2020\|EPI_ISL_698 | ···················· | ···················· | ···················· | ···················· | ···················· | ···················· |
| W | 6703 hCoV-19/United Arab Emirates/0245/2020\|EPI_ISL_698 | ···················· | ···················· | ···················· | ···················· | ···················· | ···················· |
| W | 6704 hCoV-19/United Arab Emirates/0246/2020\|EPI_ISL_698 | ···················· | ···················· | ···················· | ···················· | ···················· | ···················· |
| W | 6705 hCoV-19/United Arab Emirates/0247/2020\|EPI_ISL_698 | ···················· | ···················· | ···················· | ···················· | ···················· | ···················· |
| W | 6706 hCoV-19/United Arab Emirates/0248/2020\|EPI_ISL_698 | ···················· | ···················· | ···················· | ···················· | ···················· | ···················· |
| W | 6707 hCoV-19/United Arab Emirates/0249/2020\|EPI_ISL_698 | ···················· | ···················· | ···················· | ···················· | ···················· | ···················· |
| W | 6708 hCoV-19/United Arab Emirates/0250/2020\|EPI_ISL_698 | ···················· | ···················· | ···················· | ···················· | ···················· | ···················· |
| W | 6709 hCoV-19/United Arab Emirates/0251/2020\|EPI_ISL_698 | ···················· | ···················· | ···················· | ···················· | ···················· | ···················· |
| W | 6710 hCoV-19/United Arab Emirates/0253/2020\|EPI_ISL_698 | ···················· | ···················· | ···················· | ···················· | ···················· | ···················· |
| W | 6711 hCoV-19/United Arab Emirates/0254/2020\|EPI_ISL_698 | ···················· | ···················· | ···················· | ···················· | ···················· | ···················· |
| W | 6712 hCoV-19/India/MH-ACTREC-044/2020\|EPI_ISL_699700\|20 | ···················· | ···················· | ···················· | ···················· | ···················· | ···················· |
| W | 6713 hCoV-19/India/MH-ACTREC-045/2020\|EPI_ISL_699701\|20 | ···················· | ···················· | ···················· | ···················· | ···················· | ···················· |
| W | 6714 hCoV-19/India/MH-ACTREC-046/2020\|EPI_ISL_699702\|20 | ···················· | ···················· | ···················· | ···················· | ···················· | ···················· |
| W | 6715 hCoV-19/India/MH-ACTREC-047/2020\|EPI_ISL_699703\|20 | ···················· | ···················· | ···················· | ···················· | ···················· | ···················· |
| W | 6716 hCoV-19/India/MH-ACTREC-048/2020\|EPI_ISL_699704\|20 | ···················· | ···················· | ···················· | ···················· | ···················· | ···················· |
| W | 6719 hCoV-19/India/MH-ACTREC-052/2020\|EPI_ISL_699708\|20 | ···················· | ···················· | ···················· | ···················· | ···················· | ···················· |
| W | 6720 hCoV-19/India/MH-ACTREC-053/2020\|EPI_ISL_699709\|20 | ···················· | ···················· | ···················· | ···················· | ···················· | ···················· |
| W | 6722 hCoV-19/India/MH-ACTREC-056/2020\|EPI_ISL_699712\|20 | ···················· | ···················· | ···················· | ···················· | ···················· | ···················· |
| W | 6723 hCoV-19/India/MH-ACTREC-057/2020\|EPI_ISL_699713\|20 | ···················· | ···················· | ···················· | ···················· | ···················· | ···················· |
| W | 6724 hCoV-19/India/MH-ACTREC-058/2020\|EPI_ISL_699714\|20 | ···················· | ···················· | ···················· | ···················· | ···················· | ···················· |
| W | 6725 hCoV-19/India/MH-ACTREC-059/2020\|EPI_ISL_699715\|20 | ···················· | ···················· | ···················· | ···················· | ···················· | ···················· |
| W | 6726 hCoV-19/India/MH-ACTREC-060/2020\|EPI_ISL_699716\|20 | ···················· | ···················· | ···················· | ···················· | ···················· | ···················· |
| W | 6727 hCoV-19/India/MH-ACTREC-061/2020\|EPI_ISL_699717\|20 | ···················· | ···················· | ···················· | ···················· | ···················· | ···················· |
| W | 6728 hCoV-19/India/MH-ACTREC-062/2020\|EPI_ISL_699718\|20 | ···················· | ···················· | ···················· | ···················· | ···················· | ···················· |
| W | 6729 hCoV-19/India/MH-ACTREC-063/2020\|EPI_ISL_699719\|20 | ···················· | ···················· | ···················· | ···················· | ···················· | ···················· |
| W | 6730 hCoV-19/India/MH-ACTREC-064/2020\|EPI_ISL_699720\|20 | ···················· | ···················· | ···················· | ···················· | ···················· | ···················· |
| W | 6731 hCoV-19/India/MH-ACTREC-065/2020\|EPI_ISL_699721\|20 | ···················· | ···················· | ···················· | ···················· | ···················· | ···················· |
| W | 6733 hCoV-19/India/MH-ACTREC-068/2020\|EPI_ISL_699724\|20 | ···················· | ···················· | ···················· | ···················· | ···················· | ···················· |
| W | 6734 hCoV-19/India/MH-ACTREC-069/2020\|EPI_ISL_699725\|20 | ···················· | ···················· | ···················· | ···················· | ···················· | ···················· |
| W | 6735 hCoV-19/India/MH-ACTREC-070/2020\|EPI_ISL_699726\|20 | ···················· | ···················· | ···················· | ···················· | ···················· | ···················· |
| W | 6736 hCoV-19/India/MH-ACTREC-071/2020\|EPI_ISL_699727\|20 | ···················· | ···················· | ···················· | ···················· | ···················· | ···················· |
| W | 6737 hCoV-19/India/MH-ACTREC-072/2020\|EPI_ISL_699728\|20 | ···················· | ···················· | ···················· | ···················· | ···················· | ···················· |
| W | 6738 hCoV-19/India/MH-ACTREC-073/2020\|EPI_ISL_699729\|20 | ···················· | ···················· | ···················· | ···················· | ···················· | ···················· |
| W | 6739 hCoV-19/India/MH-ACTREC-074/2020\|EPI_ISL_699730\|20 | ···················· | ···················· | ···················· | ···················· | ···················· | ···················· |
| W | 6740 hCoV-19/India/MH-ACTREC-075/2020\|EPI_ISL_699731\|20 | ···················· | ···················· | ···················· | ···················· | ···················· | ···················· |
| W | 6741 hCoV-19/India/WB-ACTREC-076/2020\|EPI_ISL_699732\|20 | ···················· | ···················· | ···················· | ···················· | ···················· | ···················· |
| W | 6743 hCoV-19/India/MH-ACTREC-078/2020\|EPI_ISL_699734\|20 | ···················· | ···················· | ···················· | ···················· | ···················· | ···················· |
| W | 6744 hCoV-19/India/MH-ACTREC-079/2020\|EPI_ISL_699735\|20 | ···················· | ···················· | ···················· | ···················· | ···················· | ···················· |
| W | 6745 hCoV-19/India/MH-ACTREC-080/2020\|EPI_ISL_699736\|20 | ···················· | ···················· | ···················· | ···················· | ···················· | ···················· |
| W | 6746 hCoV-19/India/MH-ACTREC-081/2020\|EPI_ISL_699737\|20 | ···················· | ···················· | ···················· | ···················· | ···················· | ···················· |
| W | 6747 hCoV-19/India/MH-ACTREC-082/2020\|EPI_ISL_699738\|20 | ···················· | ···················· | ···················· | ···················· | ···················· | ···················· |
| W | 6748 hCoV-19/India/MH-ACTREC-083/2020\|EPI_ISL_699739\|20 | ···················· | ···················· | ···················· | ···················· | ···················· | ···················· |
| W | 6750 hCoV-19/India/MH-ACTREC-085/2020\|EPI_ISL_699741\|20 | ···················· | ···················· | ···················· | ···················· | ···················· | ···················· |
| W | 6751 hCoV-19/India/MH-ACTREC-086/2020\|EPI_ISL_699742\|20 | ···················· | ···················· | ···················· | ···················· | ···················· | ···················· |
| W | 6752 hCoV-19/India/MH-ACTREC-087/2020\|EPI_ISL_699743\|20 | ···················· | ···················· | ···················· | ···················· | ···················· | ···················· |
| W | 6753 hCoV-19/India/MH-ACTREC-088/2020\|EPI_ISL_699744\|20 | ···················· | ···················· | ···················· | ···················· | ···················· | ···················· |
| W | 6754 hCoV-19/India/MH-ACTREC-089/2020\|EPI_ISL_699745\|20 | ···················· | ···················· | ···················· | ···················· | ···················· | ···················· |
| W | 6755 hCoV-19/India/MH-ACTREC-090/2020\|EPI_ISL_699746\|20 | ···················· | ···················· | ···················· | ···················· | ···················· | ···················· |
| W | 6756 hCoV-19/India/MH-ACTREC-091/2020\|EPI_ISL_699747\|20 | ···················· | ···················· | ···················· | ···················· | ···················· | ···················· |
| W | 6757 hCoV-19/India/MH-ACTREC-092/2020\|EPI_ISL_699748\|20 | ···················· | ···················· | ···················· | ···················· | ···················· | ···················· |
| W | 6758 hCoV-19/India/MH-ACTREC-093/2020\|EPI_ISL_699749\|20 | ···················· | ···················· | ···················· | ···················· | ···················· | ···················· |
| W | 6759 hCoV-19/India/MH-ACTREC-094/2020\|EPI_ISL_699750\|20 | ···················· | ···················· | ···················· | ···················· | ···················· | ···················· |
| W | 6760 hCoV-19/India/MH-ACTREC-095/2020\|EPI_ISL_699751\|20 | ···T················ | ···················· | ···················· | ···················· | ···················· | ···················· |
| W | 6761 hCoV-19/India/MH-ACTREC-098/2020\|EPI_ISL_699754\|20 | ···················· | ···················· | ···················· | ···················· | ···················· | ···················· |
| W | 6762 hCoV-19/India/MH-ACTREC-099/2020\|EPI_ISL_699755\|20 | ···················· | ···················· | ···················· | ···················· | ···················· | ···················· |
| W | 6763 hCoV-19/India/MH-ACTREC-100/2020\|EPI_ISL_699756\|20 | ···················· | ···················· | ···················· | ···················· | ···················· | ···················· |
| W | 6764 hCoV-19/India/MH-ACTREC-101/2020\|EPI_ISL_699757\|20 | ···················· | ···················· | ···················· | ···················· | ···················· | ···················· |
| W | 6765 hCoV-19/India/MH-ACTREC-102/2020\|EPI_ISL_699758\|20 | ···················· | ···················· | ···················· | ···················· | ···················· | ···················· |
| W | 6766 hCoV-19/India/MH-ACTREC-103/2020\|EPI_ISL_699759\|20 | ···················· | ···················· | ···················· | ···················· | ···················· | ···················· |

| | | | | | | | | |
|---|---|---|---|---|---|---|---|---|
| W | 6767 | hCoV-19/India/MH-ACTREC-104/2020\|EPI_ISL_699760\|20 | ···················· | ···················· | ···················· | ···················· | ···················· | ···················· |
| W | 6769 | hCoV-19/India/MH-ACTREC-106/2020\|EPI_ISL_699762\|20 | ···················· | ···················· | ···················· | ···················· | ···················· | ···················· |
| W | 6770 | hCoV-19/India/MH-ACTREC-107/2020\|EPI_ISL_699763\|20 | ···················· | ···················· | ···················· | ···················· | ···················· | ···················· |
| W | 6771 | hCoV-19/India/MH-ACTREC-108/2020\|EPI_ISL_699764\|20 | ···················· | ···················· | ···················· | ···················· | ···················· | ···················· |
| W | 6772 | hCoV-19/India/MH-ACTREC-109/2020\|EPI_ISL_699765\|20 | ···················· | ···················· | ···················· | ···················· | ···················· | ···················· |
| W | 6773 | hCoV-19/India/MH-ACTREC-110/2020\|EPI_ISL_699766\|20 | ···················· | ···················· | ···················· | ···················· | ···················· | ···················· |
| W | 6774 | hCoV-19/England/ALDP-BB8F3C/2020\|EPI_ISL_703022\|20 | ···················· | ···················· | ···················· | ···················· | ···················· | ···················· |
| W | 6775 | hCoV-19/England/ALDP-BB28A0/2020\|EPI_ISL_703027\|20 | ···················· | ···················· | ···················· | ···················· | ···················· | ···················· |
| W | 6776 | hCoV-19/England/ALDP-BB3283/2020\|EPI_ISL_703028\|20 | ···················· | ···················· | ···················· | ···················· | ···················· | ···················· |
| W | 6777 | hCoV-19/England/ALDP-BB2E17/2020\|EPI_ISL_703030\|20 | ···················· | ···················· | ···················· | ···················· | ···················· | ···················· |
| W | 6778 | hCoV-19/England/ALDP-BB8BFC/2020\|EPI_ISL_703031\|20 | ···················· | ···················· | ···················· | ···················· | ···················· | ···················· |
| W | 6779 | hCoV-19/England/ALDP-BB36A5/2020\|EPI_ISL_703035\|20 | ···················· | ···················· | ···················· | ···················· | ···················· | ···················· |
| W | 6780 | hCoV-19/England/ALDP-BB9979/2020\|EPI_ISL_703037\|20 | ···················· | ·········A·········· | ···················· | ···················· | ···················· | ···················· |
| W | 6781 | hCoV-19/England/ALDP-BB8C71/2020\|EPI_ISL_703039\|20 | ···················· | ···················· | ···················· | ···················· | ···················· | ···················· |
| W | 6782 | hCoV-19/England/ALDP-BB987C/2020\|EPI_ISL_703041\|20 | ···················· | ···················· | ···················· | ···················· | ···················· | ···················· |
| W | 6783 | hCoV-19/England/ALDP-BBACD9/2020\|EPI_ISL_703042\|20 | ···················· | ···················· | ···················· | ···················· | ···················· | ···················· |
| W | 6784 | hCoV-19/England/ALDP-BB2DED/2020\|EPI_ISL_703044\|20 | ···················· | ···················· | ···················· | ···················· | ···················· | ···················· |
| W | 6785 | hCoV-19/England/ALDP-BB944B/2020\|EPI_ISL_703047\|20 | ···················· | ···················· | ···················· | ···················· | ···················· | ···················· |
| W | 6786 | hCoV-19/England/ALDP-BB3520/2020\|EPI_ISL_703049\|20 | ···················· | ···················· | ···················· | ···················· | ···················· | ···················· |
| W | 6787 | hCoV-19/England/ALDP-BB8F1E/2020\|EPI_ISL_703050\|20 | ···················· | ···················· | ···················· | ···················· | ···················· | ···················· |
| W | 6788 | hCoV-19/England/ALDP-BB5588/2020\|EPI_ISL_703052\|20 | ···················· | ···················· | ···················· | ···················· | ···················· | ···A················ |
| W | 6789 | hCoV-19/England/ALDP-BB32ED/2020\|EPI_ISL_703058\|20 | ···················· | ···················· | ···················· | ···················· | ···················· | ···················· |
| W | 6790 | hCoV-19/England/ALDP-BB8F2D/2020\|EPI_ISL_703063\|20 | ···················· | ···················· | ···················· | ···················· | ···················· | ···················· |
| W | 6791 | hCoV-19/England/ALDP-BB8F5A/2020\|EPI_ISL_703065\|20 | ···················· | ···················· | ···················· | ···················· | ···················· | ···················· |
| W | 6792 | hCoV-19/England/ALDP-BB27B2/2020\|EPI_ISL_703068\|20 | ···················· | ···················· | ···················· | ···················· | ···················· | ···················· |
| W | 6793 | hCoV-19/England/ALDP-BB55A6/2020\|EPI_ISL_703070\|20 | ···················· | ···················· | ···················· | ···················· | ···················· | ···················· |
| W | 6794 | hCoV-19/England/ALDP-BB5DDB/2020\|EPI_ISL_703075\|20 | ···················· | ···················· | ···················· | ···················· | ···················· | ···················· |
| W | 6795 | hCoV-19/England/ALDP-BB57CE/2020\|EPI_ISL_703078\|20 | ···················· | ···················· | ···················· | ···················· | ···················· | ···················· |
| W | 6796 | hCoV-19/England/ALDP-BB941E/2020\|EPI_ISL_703079\|20 | ···················· | ···················· | ···················· | ···················· | ···················· | ···················· |
| W | 6797 | hCoV-19/England/ALDP-BB56EF/2020\|EPI_ISL_703083\|20 | ···················· | ···················· | ···················· | ···················· | ···················· | ···················· |
| W | 6798 | hCoV-19/England/ALDP-BB99C4/2020\|EPI_ISL_703084\|20 | ···················· | ···················· | ···················· | ···················· | ···················· | ···················· |
| W | 6799 | hCoV-19/England/ALDP-BB2AC8/2020\|EPI_ISL_703086\|20 | ···················· | ···················· | ···················· | ···················· | ···················· | ···················· |
| W | 6800 | hCoV-19/England/ALDP-BB2B89/2020\|EPI_ISL_703091\|20 | ···················· | ···················· | ···················· | ···················· | ···A················ | ···················· |
| W | 6801 | hCoV-19/England/ALDP-BB9821/2020\|EPI_ISL_703092\|20 | ···················· | ···················· | ···················· | ···················· | ···················· | ···················· |
| W | 6802 | hCoV-19/England/ALDP-BB2925/2020\|EPI_ISL_703094\|20 | ···················· | ···················· | ···················· | ···················· | ···················· | ···················· |
| W | 6803 | hCoV-19/England/ALDP-BB996A/2020\|EPI_ISL_703095\|20 | ···················· | ···················· | ···················· | ···················· | ···················· | ···················· |
| W | 6804 | hCoV-19/England/ALDP-BB5940/2020\|EPI_ISL_703097\|20 | ···················· | ···················· | ···················· | ···················· | ···················· | ···················· |
| W | 6805 | hCoV-19/England/ALDP-BB8DF6/2020\|EPI_ISL_703098\|20 | ···················· | ···················· | ···················· | ···················· | ···················· | ···················· |
| W | 6806 | hCoV-19/England/QEUH-BCDD85/2020\|EPI_ISL_704319\|20 | ···················· | ···················· | ···················· | ···················· | ···················· | ···················· |
| W | 6807 | hCoV-19/England/QEUH-BCE4BC/2020\|EPI_ISL_704321\|20 | ···················· | ···················· | ···················· | ···················· | ···················· | ···················· |
| W | 6808 | hCoV-19/England/QEUH-BCBBE7/2020\|EPI_ISL_704323\|20 | ···················· | ···················· | ···················· | ···················· | ···················· | ···················· |
| W | 6809 | hCoV-19/England/QEUH-BCDD76/2020\|EPI_ISL_704324\|20 | ···················· | ···················· | ···················· | ···················· | ···················· | ···················· |
| W | 6810 | hCoV-19/England/QEUH-BCBAEA/2020\|EPI_ISL_704326\|20 | ···················· | ···················· | ···················· | ···················· | ···················· | ···················· |
| W | 6811 | hCoV-19/England/QEUH-BCCCC5/2020\|EPI_ISL_704327\|20 | ···················· | ···················· | ···················· | ···················· | ···················· | ···················· |
| W | 6812 | hCoV-19/England/QEUH-BCB534/2020\|EPI_ISL_704351\|20 | ···················· | ···················· | ···················· | ···················· | ···················· | ···················· |
| W | 6813 | hCoV-19/England/QEUH-BCBDE1/2020\|EPI_ISL_704352\|20 | ···················· | ···················· | ···················· | ···················· | ···················· | ···················· |
| W | 6814 | hCoV-19/England/QEUH-BCE908/2020\|EPI_ISL_704355\|20 | ···················· | ···················· | ···················· | ···················· | ···················· | ···················· |
| W | 6815 | hCoV-19/England/QEUH-BCBA9F/2020\|EPI_ISL_704357\|20 | ···················· | ···················· | ···················· | ···················· | ···················· | ···················· |
| W | 6816 | hCoV-19/England/QEUH-BCCE38/2020\|EPI_ISL_704358\|20 | ···················· | ·A·················· | ···················· | ···················· | ···················· | ···················· |
| W | 6817 | hCoV-19/England/QEUH-BCD4DB/2020\|EPI_ISL_704360\|20 | ···················· | ···················· | ···················· | ···················· | ···················· | ···················· |
| W | 6818 | hCoV-19/England/QEUH-BCB58F/2020\|EPI_ISL_704362\|20 | ···················· | ···················· | ···················· | ···················· | ···················· | ···················· |
| W | 6819 | hCoV-19/England/QEUH-BCB5E9/2020\|EPI_ISL_704363\|20 | ···················· | ·A·················· | ···················· | ···················· | ···················· | ···A················ |
| W | 6820 | hCoV-19/England/QEUH-BCC15D/2020\|EPI_ISL_704365\|20 | ···················· | ···················· | ···················· | ···················· | ···················· | ···················· |
| W | 6821 | hCoV-19/England/QEUH-BCE4F8/2020\|EPI_ISL_704368\|20 | ···················· | ···················· | ···················· | ···················· | ···················· | ···················· |
| W | 6822 | hCoV-19/England/QEUH-BCD374/2020\|EPI_ISL_704370\|20 | ···················· | ···················· | ···················· | ···················· | ···················· | ···················· |
| W | 6823 | hCoV-19/England/QEUH-BCE1B5/2020\|EPI_ISL_704376\|20 | ···················· | ···················· | ···················· | ···················· | ···················· | ···················· |
| W | 6824 | hCoV-19/England/QEUH-BCCE0B/2020\|EPI_ISL_704380\|20 | ···················· | ···················· | ···················· | ···················· | ···················· | ···················· |
| W | 6825 | hCoV-19/England/QEUH-BCDD49/2020\|EPI_ISL_704381\|20 | ···················· | ···················· | ···················· | ···················· | ···················· | ···················· |
| W | 6826 | hCoV-19/England/QEUH-BCC57F/2020\|EPI_ISL_704383\|20 | ···················· | ···················· | ···················· | ···················· | ···················· | ···················· |
| W | 6827 | hCoV-19/England/QEUH-BCCFCC/2020\|EPI_ISL_704386\|20 | ···················· | ···················· | ···················· | ···················· | ···················· | ···················· |
| W | 6828 | hCoV-19/England/QEUH-BCB5DA/2020\|EPI_ISL_704388\|20 | ···················· | ···················· | ···················· | ···················· | ···················· | ···················· |
| W | 6829 | hCoV-19/England/QEUH-BCDDEF/2020\|EPI_ISL_704391\|20 | ···················· | ···················· | ···················· | ···················· | ···················· | ···················· |
| W | 6830 | hCoV-19/England/QEUH-BCE22B/2020\|EPI_ISL_704394\|20 | ···················· | ···················· | ···················· | ···················· | ···················· | ···················· |
| W | 6831 | hCoV-19/England/QEUH-BCCAE9/2020\|EPI_ISL_704396\|20 | ···················· | ···················· | ···················· | ···················· | ···················· | ···················· |
| W | 6832 | hCoV-19/England/QEUH-BCB59E/2020\|EPI_ISL_704399\|20 | ···················· | ···················· | ···················· | ···················· | ···················· | ···················· |
| W | 6833 | hCoV-19/Scotland/CVR5888/2020\|EPI_ISL_705648\|2020- | ···················· | ···················· | ···················· | ···················· | ···················· | ···················· |
| W | 6834 | hCoV-19/Scotland/CVR5890/2020\|EPI_ISL_705649\|2020- | ···················· | ···················· | ···················· | ···················· | ···················· | ···················· |
| W | 6835 | hCoV-19/Scotland/CVR5891/2020\|EPI_ISL_705650\|2020- | ···················· | ···················· | ···················· | ···················· | ···················· | ···················· |
| W | 6836 | hCoV-19/Scotland/CVR5892/2020\|EPI_ISL_705651\|2020- | ···················· | ···················· | ···················· | ···················· | ···················· | ···················· |
| W | 6837 | hCoV-19/Scotland/CVR5893/2020\|EPI_ISL_705652\|2020- | ···················· | ···················· | ···················· | ···················· | ···················· | ···················· |
| W | 6838 | hCoV-19/Scotland/CVR5894/2020\|EPI_ISL_705653\|2020- | ···················· | ···················· | ···················· | ···················· | ···················· | ···················· |
| W | 6839 | hCoV-19/Scotland/CVR5895/2020\|EPI_ISL_705654\|2020- | ···················· | ···················· | ···················· | ···················· | ···················· | ···················· |
| W | 6840 | hCoV-19/Scotland/CVR5896/2020\|EPI_ISL_705655\|2020- | ···················· | ···················· | ···················· | ···················· | ···················· | ···················· |
| W | 6841 | hCoV-19/Scotland/CVR5897/2020\|EPI_ISL_705656\|2020- | ···················· | ···················· | ···················· | ···················· | ···················· | ···················· |
| W | 6842 | hCoV-19/Scotland/CVR5898/2020\|EPI_ISL_705657\|2020- | ···················· | ···················· | ···················· | ···················· | ···················· | ···················· |
| W | 6843 | hCoV-19/Scotland/CVR5900/2020\|EPI_ISL_705658\|2020- | ···················· | ···················· | ···················· | ···················· | ···················· | ···················· |
| W | 6844 | hCoV-19/Scotland/CVR5901/2020\|EPI_ISL_705659\|2020- | ···················· | ···················· | ···················· | ···················· | ···················· | ···················· |
| W | 6845 | hCoV-19/Scotland/CVR5935/2020\|EPI_ISL_705680\|2020- | ···················· | ···················· | ···················· | ···················· | ···················· | ···················· |
| W | 6846 | hCoV-19/Scotland/CVR5937/2020\|EPI_ISL_705681\|2020- | ···················· | ···················· | ···················· | ···················· | ···················· | ···················· |
| W | 6847 | hCoV-19/Scotland/CVR5940/2020\|EPI_ISL_705682\|2020- | ···················· | ···················· | ···················· | ···················· | ···················· | ···················· |

| | | | | | | | | |
|---|---|---|---|---|---|---|---|---|
| W | 6848 | hCoV-19/Scotland/CVR5941/2020\|EPI_ISL_705683\|2020- | ···················· | ···················· | ···················· | ···················· | ···················· | ···················· |
| W | 6849 | hCoV-19/Scotland/CVR5942/2020\|EPI_ISL_705684\|2020- | ···················· | ···················· | ···················· | ···················· | ···················· | ···················· |
| W | 6850 | hCoV-19/Scotland/CVR5944/2020\|EPI_ISL_705685\|2020- | ···················· | ···················· | ···················· | ···················· | ···················· | ···················· |
| W | 6851 | hCoV-19/Scotland/CVR5945/2020\|EPI_ISL_705686\|2020- | ···················· | ···················· | ···················· | ···················· | ···················· | ···················· |
| W | 6852 | hCoV-19/Scotland/CVR5946/2020\|EPI_ISL_705687\|2020- | ···················· | ···················· | ···················· | ···················· | ···················· | ···················· |
| W | 6853 | hCoV-19/Scotland/CVR5948/2020\|EPI_ISL_705688\|2020- | ···················· | ···················· | ···················· | ···················· | ···················· | ···················· |
| W | 6854 | hCoV-19/Scotland/CVR5950/2020\|EPI_ISL_705689\|2020- | ···················· | ···················· | ···················· | ···················· | ···················· | ···················· |
| W | 6855 | hCoV-19/Scotland/CVR5951/2020\|EPI_ISL_705690\|2020- | ···················· | ···················· | ···················· | ···················· | ···················· | ···················· |
| W | 6856 | hCoV-19/Scotland/CVR5952/2020\|EPI_ISL_705691\|2020- | ···················· | ···················· | ···················· | ···················· | ···················· | ···················· |
| W | 6857 | hCoV-19/Scotland/CVR5953/2020\|EPI_ISL_705692\|2020- | ···················· | ···················· | ···················· | ···················· | ···················· | ···················· |
| W | 6858 | hCoV-19/Scotland/CVR5954/2020\|EPI_ISL_705693\|2020- | ···················· | ···················· | ···················· | ···················· | ···················· | ···················· |
| W | 6859 | hCoV-19/Scotland/CVR5955/2020\|EPI_ISL_705694\|2020- | ···················· | ···················· | ···················· | ···················· | ···················· | ···················· |
| W | 6860 | hCoV-19/Scotland/CVR5956/2020\|EPI_ISL_705695\|2020- | ···················· | ···················· | ···················· | ···················· | ···················· | ···················· |
| W | 6861 | hCoV-19/Scotland/CVR5957/2020\|EPI_ISL_705696\|2020- | ···················· | ···················· | ···················· | ···················· | ···················· | ···················· |
| W | 6862 | hCoV-19/Scotland/CVR5958/2020\|EPI_ISL_705697\|2020- | ···················· | ···················· | ···················· | ···················· | ···················· | ···················· |
| W | 6863 | hCoV-19/Scotland/CVR5961/2020\|EPI_ISL_705698\|2020- | ···················· | ···················· | ···················· | ···················· | ···················· | ···················· |
| W | 6864 | hCoV-19/Scotland/CVR5962/2020\|EPI_ISL_705699\|2020- | ···················· | ···················· | ···················· | ···················· | ···················· | ···················· |
| W | 6865 | hCoV-19/England/SHEF-10CEFE0/2020\|EPI_ISL_706980\|2 | ···················· | ···················· | ···················· | ···················· | ···················· | ···················· |
| W | 6866 | hCoV-19/England/SHEF-10D090B/2020\|EPI_ISL_706981\|2 | ···················· | ···················· | ···················· | ···················· | ···················· | ···················· |
| W | 6867 | hCoV-19/England/SHEF-10D0ABD/2020\|EPI_ISL_706982\|2 | ···················· | ···················· | ···················· | ···················· | ···················· | ···················· |
| W | 6868 | hCoV-19/England/SHEF-10D2893/2020\|EPI_ISL_706983\|2 | ···················· | ···················· | ···················· | ···················· | ···················· | ···················· |
| W | 6869 | hCoV-19/England/SHEF-10D2BD6/2020\|EPI_ISL_706984\|2 | ···················· | ···················· | ···················· | ···················· | ···················· | ···················· |
| W | 6870 | hCoV-19/England/SHEF-10D2D2B/2020\|EPI_ISL_706985\|2 | ···················· | ···················· | ···················· | ···················· | ···················· | ···················· |
| W | 6871 | hCoV-19/England/SHEF-10D3C69/2020\|EPI_ISL_706986\|2 | ···················· | ···················· | ···················· | ···················· | ···················· | ···················· |
| W | 6872 | hCoV-19/England/SHEF-10D4451/2020\|EPI_ISL_706987\|2 | ···················· | ···················· | ···················· | ···················· | ···················· | ···················· |
| W | 6873 | hCoV-19/England/SHEF-10D6A11/2020\|EPI_ISL_706988\|2 | ···················· | ···················· | ···················· | ···················· | ···················· | ···················· |
| W | 6874 | hCoV-19/England/SHEF-10D6CC0/2020\|EPI_ISL_706989\|2 | ···················· | ···················· | ···················· | ···················· | ···················· | ···················· |
| W | 6875 | hCoV-19/England/CAMC-BFBEF4/2020\|EPI_ISL_709200\|20 | ···················· | ···················· | ···················· | ···················· | ···················· | ···················· |
| W | 6876 | hCoV-19/England/QEUH-BF0D10/2020\|EPI_ISL_709201\|20 | ···················· | ···················· | ···················· | ···················· | ···················· | ···················· |
| W | 6877 | hCoV-19/England/CAMC-BFB911/2020\|EPI_ISL_709204\|20 | ···················· | ···················· | ···················· | ···················· | ···················· | ···················· |
| W | 6878 | hCoV-19/England/CAMC-BFBF00/2020\|EPI_ISL_709206\|20 | ···················· | ···················· | ···················· | ···················· | ···················· | ···················· |
| W | 6879 | hCoV-19/England/CAMC-BFB4A7/2020\|EPI_ISL_709207\|20 | ···················· | ···················· | ···················· | ···················· | ···················· | ···················· |
| W | 6880 | hCoV-19/England/CAMC-BFB29E/2020\|EPI_ISL_709209\|20 | ···················· | ···················· | ···················· | ···················· | ···················· | ···················· |
| W | 6881 | hCoV-19/England/CAMC-BF480C/2020\|EPI_ISL_709220\|20 | ···················· | ···················· | ···················· | ···················· | ·····T·············· | ···················· |
| W | 6882 | hCoV-19/England/CAMC-BFBC90/2020\|EPI_ISL_709224\|20 | ···················· | ···················· | ···················· | ···················· | ···················· | ···················· |
| W | 6883 | hCoV-19/England/QEUH-BF0D3E/2020\|EPI_ISL_709231\|20 | ···················· | ···················· | ···················· | ···················· | ···················· | ···················· |
| W | 6884 | hCoV-19/England/CAMC-BFBA0F/2020\|EPI_ISL_709232\|20 | ···················· | ···················· | ···················· | ···················· | ···················· | ···················· |
| W | 6885 | hCoV-19/England/CAMC-BFBC54/2020\|EPI_ISL_709233\|20 | ···················· | ···················· | ···················· | ···················· | ·····T·············· | ···················· |
| W | 6886 | hCoV-19/England/QEUH-BF0A55/2020\|EPI_ISL_709234\|20 | ···················· | ···················· | ···················· | ···················· | ···················· | ···················· |
| W | 6887 | hCoV-19/England/QEUH-BF09A3/2020\|EPI_ISL_709236\|20 | ···················· | ···················· | ···················· | ···················· | ···················· | ···················· |
| W | 6888 | hCoV-19/England/CAMC-BFBAB4/2020\|EPI_ISL_709238\|20 | ···················· | ···················· | ···················· | ···················· | ···················· | ···················· |
| W | 6889 | hCoV-19/England/QEUH-BF0CAA/2020\|EPI_ISL_709239\|20 | ···················· | ···················· | ···················· | ···················· | ···················· | ···················· |
| W | 6891 | hCoV-19/USA/CO-CDPHE-2004230209/2020\|EPI_ISL_71020 | ···················· | ···················· | ···················· | ···················· | ···················· | ···················· |
| W | 6892 | hCoV-19/USA/CO-CDPHE-2004230217/2020\|EPI_ISL_71020 | ···················· | ···················· | ···················· | ···················· | ···················· | ···················· |
| W | 6896 | hCoV-19/USA/CO-CDPHE-2005110165/2020\|EPI_ISL_71021 | ···················· | ···················· | ···················· | ···················· | ···················· | ···················· |
| W | 6897 | hCoV-19/USA/CO-CDPHE-2005220143/2020\|EPI_ISL_71021 | ···················· | ···················· | ···················· | ···················· | ···················· | ···················· |
| W | 6898 | hCoV-19/USA/CO-CDPHE-2006280631/2020\|EPI_ISL_71021 | ···················· | ···················· | ···················· | ···················· | ···················· | ···················· |
| W | 6899 | hCoV-19/USA/CO-CDPHE-2007230810/2020\|EPI_ISL_71021 | ···················· | ···················· | ···················· | ···················· | ···················· | ···················· |
| W | 6900 | hCoV-19/USA/CO-CDPHE-2007242803/2020\|EPI_ISL_71021 | ···················· | ···················· | ···················· | ···················· | ···················· | ···················· |
| W | 6901 | hCoV-19/USA/CO-CDPHE-2007250713/2020\|EPI_ISL_71021 | ···················· | ···················· | ···················· | ···················· | ···················· | ···················· |
| W | 6902 | hCoV-19/USA/CO-CDPHE-2008161331/2020\|EPI_ISL_71021 | ···················· | ···················· | ···················· | ···················· | ···················· | ···················· |
| W | 6903 | hCoV-19/USA/CO-CDPHE-2008180319/2020\|EPI_ISL_71021 | ···················· | ···················· | ···················· | ···················· | ···················· | ···················· |
| W | 6904 | hCoV-19/USA/CO-CDPHE-2008180407/2020\|EPI_ISL_71021 | ···················· | ···················· | ···················· | ···················· | ···················· | ···················· |
| W | 6905 | hCoV-19/USA/CO-CDPHE-2008181078/2020\|EPI_ISL_71022 | ···················· | ···················· | ···················· | ···················· | ···················· | ···················· |
| W | 6906 | hCoV-19/USA/CO-CDPHE-2008181628/2020\|EPI_ISL_71022 | ···················· | ···················· | ···················· | ···················· | ···················· | ···················· |
| W | 6907 | hCoV-19/USA/CO-CDPHE-2009042396/2020\|EPI_ISL_71022 | ···················· | ·A·················· | ···················· | ···················· | ···················· | ···················· |
| W | 6908 | hCoV-19/USA/CO-CDPHE-2009053317/2020\|EPI_ISL_71022 | ···················· | ···················· | ···················· | ···················· | ···················· | ···················· |
| W | 6909 | hCoV-19/USA/CO-CDPHE-2009131675/2020\|EPI_ISL_71022 | ···················· | ·A·················· | ···················· | ···················· | ···················· | ···················· |
| W | 6910 | hCoV-19/USA/CO-CDPHE-2009131687/2020\|EPI_ISL_71022 | ···················· | ·A·················· | ···················· | ···················· | ···················· | ···················· |
| W | 6911 | hCoV-19/USA/CO-CDPHE-2009131689/2020\|EPI_ISL_71022 | ···················· | ·A·················· | ···················· | ···················· | ···················· | ···················· |
| W | 6912 | hCoV-19/USA/CO-CDPHE-2009131699/2020\|EPI_ISL_71022 | ···················· | ·A·················· | ···················· | ···················· | ···················· | ···················· |
| W | 6913 | hCoV-19/USA/AZ-TG397126/2020\|EPI_ISL_694845\|2020-0 | ···················· | ···················· | ···················· | ···················· | ···················· | ···················· |
| W | 6914 | hCoV-19/USA/AZ-TG397199/2020\|EPI_ISL_694846\|2020-0 | ···················· | ···················· | ···················· | ···················· | ···················· | ···················· |
| W | 6915 | hCoV-19/USA/AZ-TG398427/2020\|EPI_ISL_694847\|2020-0 | ···················· | ···················· | ···················· | ···················· | ···················· | ···················· |
| W | 6916 | hCoV-19/USA/AZ-TG398623/2020\|EPI_ISL_694849\|2020-0 | ···················· | ·A·················· | ···················· | ···················· | ···················· | ···················· |
| W | 6917 | hCoV-19/USA/AZ-TG398670/2020\|EPI_ISL_694850\|2020-0 | ···················· | ···················· | ···················· | ···················· | ···················· | ···················· |
| W | 6918 | hCoV-19/USA/AZ-TG402010/2020\|EPI_ISL_694855\|2020-0 | ···················· | ···················· | ···················· | ···················· | ···················· | ···················· |
| W | 6919 | hCoV-19/USA/AZ-TG402310/2020\|EPI_ISL_694856\|2020-0 | ···················· | ···················· | ···················· | ···················· | ···················· | ···················· |
| W | 6920 | hCoV-19/USA/AZ-TG402493/2020\|EPI_ISL_694857\|2020-0 | ···················· | ···················· | ···················· | ···················· | ···················· | ···················· |
| W | 6921 | hCoV-19/USA/AZ-TG402761/2020\|EPI_ISL_694858\|2020-0 | ···················· | ···················· | ···················· | ···················· | ···················· | ···················· |
| W | 6922 | hCoV-19/USA/AZ-TG402846/2020\|EPI_ISL_694859\|2020-0 | ···················· | ···················· | ···················· | ···················· | ···················· | ···················· |
| W | 6923 | hCoV-19/USA/AZ-TG403691/2020\|EPI_ISL_694864\|2020-0 | ···················· | ·A·················· | ···················· | ···················· | ···················· | ···················· |
| W | 6924 | hCoV-19/USA/AZ-TG403895/2020\|EPI_ISL_694865\|2020-0 | ···················· | ·A·················· | ···················· | ···················· | ···················· | ···················· |
| W | 6925 | hCoV-19/USA/AZ-TG404376/2020\|EPI_ISL_694866\|2020-0 | ···················· | ·A·················· | ···················· | ···················· | ···················· | ···················· |
| W | 6926 | hCoV-19/USA/AZ-TG404882/2020\|EPI_ISL_694867\|2020-0 | ···················· | ·A·················· | ···················· | ···················· | ···················· | ···················· |
| W | 6927 | hCoV-19/USA/AZ-TG405021/2020\|EPI_ISL_694868\|2020-0 | ···················· | ···················· | ···················· | ···················· | ···················· | ···················· |
| W | 6928 | hCoV-19/USA/AZ-TG405037/2020\|EPI_ISL_694870\|2020-0 | ···················· | ···················· | ···················· | ···················· | ···················· | ···················· |
| W | 6929 | hCoV-19/USA/AZ-TG405412/2020\|EPI_ISL_694871\|2020-0 | ···················· | ·A·················· | ···················· | ···················· | ···················· | ···················· |
| W | 6930 | hCoV-19/United Arab Emirates/0125/2020\|EPI_ISL_698 | ···················· | ···················· | ···················· | ···················· | ···················· | ···················· |
| W | 6931 | hCoV-19/United Arab Emirates/0126/2020\|EPI_ISL_698 | ···················· | ···················· | ···················· | ···················· | ···················· | ···················· |

| | | | | | | | | |
|---|---|---|---|---|---|---|---|---|
| W | 6932 | hCoV-19/United Arab Emirates/0127/2020\|EPI_ISL_698 | .................... | .................... | .................... | .................... | .................... | .................... |
| W | 6933 | hCoV-19/United Arab Emirates/0128/2020\|EPI_ISL_698 | .................... | .................... | .................... | .................... | .................... | .................... |
| W | 6934 | hCoV-19/United Arab Emirates/0129/2020\|EPI_ISL_698 | .................... | .................... | .................... | .................... | .................... | .................... |
| W | 6935 | hCoV-19/United Arab Emirates/0130/2020\|EPI_ISL_698 | .................... | .................... | .................... | .................... | .................... | .................... |
| W | 6936 | hCoV-19/United Arab Emirates/0131/2020\|EPI_ISL_698 | .................... | .................... | .................... | .................... | .................... | .................... |
| W | 6937 | hCoV-19/United Arab Emirates/0132/2020\|EPI_ISL_698 | .................... | .................... | .................... | .................... | .................... | .................... |
| W | 6938 | hCoV-19/United Arab Emirates/0136/2020\|EPI_ISL_698 | .................... | .................... | .................... | .................... | .................... | .................... |
| W | 6939 | hCoV-19/United Arab Emirates/0137/2020\|EPI_ISL_698 | .................... | .................... | .................... | .................... | .................... | .................... |
| W | 6940 | hCoV-19/United Arab Emirates/0138/2020\|EPI_ISL_698 | .................... | .................... | .................... | .................... | .................... | .................... |
| W | 6941 | hCoV-19/United Arab Emirates/0139/2020\|EPI_ISL_698 | .................... | .................... | .................... | .................... | .................... | .................... |
| W | 6942 | hCoV-19/United Arab Emirates/0140/2020\|EPI_ISL_698 | .................... | .................... | .................... | .................... | .................... | .................... |
| W | 6943 | hCoV-19/United Arab Emirates/0141/2020\|EPI_ISL_698 | .................... | .................... | .................... | .................... | .................... | .................... |
| W | 6944 | hCoV-19/United Arab Emirates/0142/2020\|EPI_ISL_698 | .................... | .................... | .................... | .................... | .................... | .................... |
| W | 6945 | hCoV-19/United Arab Emirates/0143/2020\|EPI_ISL_698 | .................... | .................... | .................... | .................... | .................... | .................... |
| W | 6946 | hCoV-19/United Arab Emirates/0144/2020\|EPI_ISL_698 | .................... | .................... | .................... | .................... | .................... | .................... |
| W | 6947 | hCoV-19/United Arab Emirates/0145/2020\|EPI_ISL_698 | .................... | .................... | .................... | .................... | .................... | .................... |
| W | 6948 | hCoV-19/United Arab Emirates/0146/2020\|EPI_ISL_698 | .................... | .................... | .................... | .................... | .................... | .................... |
| W | 6949 | hCoV-19/United Arab Emirates/0147/2020\|EPI_ISL_698 | .................... | .................... | .................... | .................... | .................... | .................... |
| W | 6950 | hCoV-19/United Arab Emirates/0148/2020\|EPI_ISL_698 | .................... | .................... | .................... | .................... | .................... | .................... |
| W | 6951 | hCoV-19/United Arab Emirates/0149/2020\|EPI_ISL_698 | .................... | .................... | .................... | .................... | .................... | .................... |
| W | 6952 | hCoV-19/United Arab Emirates/0150/2020\|EPI_ISL_698 | .................... | .................... | .................... | .................... | .................... | .................... |
| W | 6953 | hCoV-19/United Arab Emirates/0151/2020\|EPI_ISL_698 | .................... | .................... | .................... | .................... | .................... | .................... |
| W | 6954 | hCoV-19/United Arab Emirates/0152/2020\|EPI_ISL_698 | .................... | .................... | .................... | .................... | .................... | .................... |
| W | 6955 | hCoV-19/United Arab Emirates/0153/2020\|EPI_ISL_698 | .................... | .................... | .................... | .................... | .................... | .................... |
| W | 6956 | hCoV-19/United Arab Emirates/0154/2020\|EPI_ISL_698 | .................... | .................... | .................... | .................... | .................... | .................... |
| W | 6957 | hCoV-19/United Arab Emirates/0155/2020\|EPI_ISL_698 | .................... | .................... | .................... | .................... | .................... | .................... |
| W | 6958 | hCoV-19/United Arab Emirates/0156/2020\|EPI_ISL_698 | .................... | .................... | .................... | .................... | .................... | .................... |
| W | 6959 | hCoV-19/United Arab Emirates/0157/2020\|EPI_ISL_698 | .................... | .................... | .................... | .................... | .................... | .................... |
| W | 6960 | hCoV-19/United Arab Emirates/0158/2020\|EPI_ISL_698 | .................... | .................... | .................... | .................... | .................... | .................... |
| W | 6961 | hCoV-19/United Arab Emirates/0159/2020\|EPI_ISL_698 | .................... | .................... | .................... | .................... | .................... | .................... |
| W | 6962 | hCoV-19/United Arab Emirates/0160/2020\|EPI_ISL_698 | .................... | .................... | .................... | .................... | .................... | .................... |
| W | 6963 | hCoV-19/United Arab Emirates/0161/2020\|EPI_ISL_698 | .................... | .................... | .................... | .................... | .................... | .................... |
| W | 6964 | hCoV-19/United Arab Emirates/0162/2020\|EPI_ISL_698 | .................... | .................... | .................... | .................... | .................... | .................... |
| W | 6965 | hCoV-19/United Arab Emirates/0163/2020\|EPI_ISL_698 | .................... | .................... | .................... | .................... | .................... | .................... |
| W | 6966 | hCoV-19/United Arab Emirates/0164/2020\|EPI_ISL_698 | .................... | .................... | .................... | .................... | .................... | .................... |
| W | 6967 | hCoV-19/United Arab Emirates/0165/2020\|EPI_ISL_698 | .................... | .................... | .................... | .................... | .................... | .................... |
| W | 6968 | hCoV-19/United Arab Emirates/0166/2020\|EPI_ISL_698 | .................... | .................... | .................... | .................... | .................... | .................... |
| W | 6969 | hCoV-19/United Arab Emirates/0167/2020\|EPI_ISL_698 | .................... | .................... | .................... | .................... | .................... | .................... |
| W | 6970 | hCoV-19/United Arab Emirates/0168/2020\|EPI_ISL_698 | .................... | .................... | .................... | .................... | .................... | .................... |
| W | 6971 | hCoV-19/United Arab Emirates/0169/2020\|EPI_ISL_698 | .................... | .................... | .................... | .................... | .................... | .................... |
| W | 6972 | hCoV-19/United Arab Emirates/0170/2020\|EPI_ISL_698 | .................... | .................... | .................... | .................... | .................... | .................... |
| W | 6973 | hCoV-19/United Arab Emirates/0171/2020\|EPI_ISL_698 | .................... | .................... | .................... | .................... | .................... | .................... |
| W | 6974 | hCoV-19/United Arab Emirates/0172/2020\|EPI_ISL_698 | .................... | .................... | .................... | .................... | .................... | .................... |
| W | 6975 | hCoV-19/United Arab Emirates/0175/2020\|EPI_ISL_698 | .................... | .................... | .................... | .................... | .................... | .................... |
| W | 6976 | hCoV-19/United Arab Emirates/0176/2020\|EPI_ISL_698 | .................... | .................... | .................... | .................... | .................... | .................... |
| W | 6977 | hCoV-19/United Arab Emirates/0177/2020\|EPI_ISL_698 | .................... | .................... | .................... | .................... | .................... | .................... |
| W | 6978 | hCoV-19/United Arab Emirates/0178/2020\|EPI_ISL_698 | .................... | .................... | .................... | .................... | .................... | .................... |
| W | 6979 | hCoV-19/United Arab Emirates/0179/2020\|EPI_ISL_698 | .................... | .................... | .................... | .................... | .................... | .................... |
| W | 6980 | hCoV-19/United Arab Emirates/0180/2020\|EPI_ISL_698 | .................... | .................... | .................... | .................... | .................... | .................... |
| W | 6981 | hCoV-19/United Arab Emirates/0181/2020\|EPI_ISL_698 | .................... | .................... | .................... | .................... | .................... | .................... |
| W | 6982 | hCoV-19/United Arab Emirates/0182/2020\|EPI_ISL_698 | .................... | .................... | .................... | .................... | .................... | .................... |
| W | 6983 | hCoV-19/Tunisia/61689/2020\|EPI_ISL_699655\|2020-11- | .................... | .................... | .................... | .................... | .................... | .................... |
| W | 6984 | hCoV-19/Tunisia/5008/2020\|EPI_ISL_699656\|2020-03-2 | .................... | .................... | .................... | .................... | .................... | .................... |
| W | 6985 | hCoV-19/Tunisia/3942/2020\|EPI_ISL_699657\|2020-03-1 | .................... | .................... | .................... | .................... | .................... | .................... |
| W | 6986 | hCoV-19/India/UP-ACTREC-001/2020\|EPI_ISL_699658\|20 | .................... | .................... | .................... | .................... | .................... | .................... |
| W | 6987 | hCoV-19/India/MH-ACTREC-002/2020\|EPI_ISL_699659\|20 | .................... | .................... | .................... | .................... | .................... | .................... |
| W | 6988 | hCoV-19/India/MH-ACTREC-003/2020\|EPI_ISL_699660\|20 | .................... | .................... | .................... | .................... | .................... | .................... |
| W | 6989 | hCoV-19/India/MH-ACTREC-004/2020\|EPI_ISL_699661\|20 | .................... | .................... | .................... | .................... | .................... | .................... |
| W | 6990 | hCoV-19/India/MH-ACTREC-005/2020\|EPI_ISL_699662\|20 | .................... | .................... | .................... | .................... | .................... | .................... |
| W | 6991 | hCoV-19/India/MH-ACTREC-006/2020\|EPI_ISL_699663\|20 | .................... | .................... | .................... | .................... | .................... | .................... |
| W | 6992 | hCoV-19/India/MH-ACTREC-007/2020\|EPI_ISL_699664\|20 | .................... | .................... | .................... | .................... | .................... | .................... |
| W | 6993 | hCoV-19/India/MH-ACTREC-008/2020\|EPI_ISL_699665\|20 | .................... | .................... | .................... | .................... | .................... | .................... |
| W | 6994 | hCoV-19/India/MH-ACTREC-009/2020\|EPI_ISL_699666\|20 | .................... | .................... | .................... | .................... | .................... | .................... |
| W | 6995 | hCoV-19/India/MH-ACTREC-010/2020\|EPI_ISL_699667\|20 | .................... | .................... | .................... | .................... | .................... | .................... |
| W | 6996 | hCoV-19/India/MH-ACTREC-011/2020\|EPI_ISL_699668\|20 | .................... | .................... | .................... | .................... | .................... | .................... |
| W | 6997 | hCoV-19/India/MH-ACTREC-013/2020\|EPI_ISL_699669\|20 | .................... | .................... | .................... | .................... | .................... | .................... |
| W | 6998 | hCoV-19/India/MH-ACTREC-014/2020\|EPI_ISL_699670\|20 | .................... | .................... | .................... | .................... | .................... | .................... |
| W | 6999 | hCoV-19/India/MH-ACTREC-015/2020\|EPI_ISL_699671\|20 | .................... | .................... | .................... | .................... | .................... | .................... |
| W | 7000 | hCoV-19/India/MH-ACTREC-016/2020\|EPI_ISL_699672\|20 | .................... | .................... | .................... | .................... | .................... | .................... |
| W | 7001 | hCoV-19/India/MH-ACTREC-017/2020\|EPI_ISL_699673\|20 | .................... | .................... | .................... | .................... | .................... | .................... |
| W | 7002 | hCoV-19/India/MH-ACTREC-018/2020\|EPI_ISL_699674\|20 | .................... | .................... | .................... | .................... | .................... | .................... |
| W | 7003 | hCoV-19/India/MH-ACTREC-019/2020\|EPI_ISL_699675\|20 | .................... | .................... | .................... | .................... | .................... | .................... |
| W | 7004 | hCoV-19/India/MH-ACTREC-020/2020\|EPI_ISL_699676\|20 | .................... | .................... | .................... | .................... | .................... | .................... |
| W | 7005 | hCoV-19/India/MH-ACTREC-021/2020\|EPI_ISL_699677\|20 | .................... | .................... | .................... | .................... | .................... | .................... |
| W | 7006 | hCoV-19/India/MH-ACTREC-022/2020\|EPI_ISL_699678\|20 | .................... | .................... | .................... | .................... | .................... | .................... |
| W | 7007 | hCoV-19/India/MH-ACTREC-023/2020\|EPI_ISL_699679\|20 | .................... | .................... | .................... | .................... | .................... | .................... |
| W | 7008 | hCoV-19/India/MH-ACTREC-024/2020\|EPI_ISL_699680\|20 | .................... | .................... | .................... | .................... | .................... | .................... |
| W | 7010 | hCoV-19/India/MH-ACTREC-026/2020\|EPI_ISL_699682\|20 | .................... | .................... | .................... | .................... | .................... | .................... |
| W | 7011 | hCoV-19/India/MH-ACTREC-027/2020\|EPI_ISL_699683\|20 | .................... | .................... | .................... | .................... | .................... | .................... |
| W | 7012 | hCoV-19/India/MH-ACTREC-028/2020\|EPI_ISL_699684\|20 | .................... | .................... | .................... | .................... | .................... | .................... |

| | | | | | | | | |
|---|---|---|---|---|---|---|---|---|
| W | 7013 hCoV-19/India/MH-ACTREC-029/2020\|EPI_ISL_699685\|20 | ···················· | ···················· | ···················· | ···················· | ···················· | ···················· | |
| W | 7014 hCoV-19/India/MH-ACTREC-030/2020\|EPI_ISL_699686\|20 | ···················· | ···················· | ···················· | ···················· | ···················· | ···················· | |
| W | 7015 hCoV-19/India/MH-ACTREC-031/2020\|EPI_ISL_699687\|20 | ···················· | ···················· | ···················· | ···················· | ···················· | ···················· | |
| W | 7016 hCoV-19/India/MH-ACTREC-032/2020\|EPI_ISL_699688\|20 | ···················· | ···················· | ···················· | ···················· | ···················· | ···················· | |
| W | 7018 hCoV-19/India/MH-ACTREC-035/2020\|EPI_ISL_699691\|20 | ···················· | ·A·················· | ···················· | ···················· | ···················· | ···················· | |
| W | 7019 hCoV-19/India/MH-ACTREC-036/2020\|EPI_ISL_699692\|20 | ···················· | ···················· | ···················· | ···················· | ···················· | ···················· | |
| W | 7020 hCoV-19/India/MH-ACTREC-037/2020\|EPI_ISL_699693\|20 | ···················· | ···················· | ···················· | ···················· | ···················· | ···················· | |
| W | 7021 hCoV-19/India/MH-ACTREC-038/2020\|EPI_ISL_699694\|20 | ···················· | ···················· | ···················· | ···················· | ···················· | ···················· | |
| W | 7022 hCoV-19/India/MH-ACTREC-039/2020\|EPI_ISL_699695\|20 | ···················· | ···················· | ···················· | ···················· | ···················· | ···················· | |
| W | 7023 hCoV-19/India/MH-ACTREC-040/2020\|EPI_ISL_699696\|20 | ···················· | ···················· | ···················· | ···················· | ···················· | ···················· | |
| W | 7024 hCoV-19/India/MH-ACTREC-041/2020\|EPI_ISL_699697\|20 | ···················· | ···················· | ···················· | ···················· | ···················· | ···················· | |
| W | 7025 hCoV-19/India/MH-ACTREC-042/2020\|EPI_ISL_699698\|20 | ···················· | ···················· | ···················· | ···················· | ···················· | ···················· | |
| W | 7026 hCoV-19/India/MH-ACTREC-043/2020\|EPI_ISL_699699\|20 | ···················· | ···················· | ···················· | ···················· | ···················· | ···················· | |
| W | 7027 hCoV-19/England/ALDP-BB598C/2020\|EPI_ISL_703120\|20 | ···················· | ···················· | ···················· | ···················· | ···················· | ···················· | |
| W | 7028 hCoV-19/England/ALDP-BB9AFE/2020\|EPI_ISL_703122\|20 | ···················· | ···················· | ···················· | ···················· | ···················· | ···················· | |
| W | 7029 hCoV-19/England/ALDP-BB57BF/2020\|EPI_ISL_703123\|20 | ···················· | ···················· | ···················· | ···················· | ···················· | ···················· | |
| W | 7030 hCoV-19/England/ALDP-BB31A4/2020\|EPI_ISL_703125\|20 | ···················· | ···················· | ···················· | ···················· | ···················· | ···················· | |
| W | 7031 hCoV-19/England/ALDP-BB9DAA/2020\|EPI_ISL_703126\|20 | ···················· | ···················· | ···················· | ···················· | ···················· | ···················· | |
| W | 7032 hCoV-19/England/ALDP-BB57DD/2020\|EPI_ISL_703128\|20 | ···················· | ···················· | ···················· | ···················· | ···················· | ···················· | |
| W | 7033 hCoV-19/England/QEUH-BCD6B7/2020\|EPI_ISL_704400\|20 | ···················· | ···················· | ···················· | ···················· | ···················· | ···················· | |
| W | 7034 hCoV-19/England/QEUH-BCB77A/2020\|EPI_ISL_704402\|20 | ···················· | ···················· | ···················· | ···················· | ···················· | ···················· | |
| W | 7035 hCoV-19/England/QEUH-BCCFAE/2020\|EPI_ISL_704404\|20 | ···················· | ···················· | ···················· | ···················· | ···················· | ···················· | |
| W | 7036 hCoV-19/England/QEUH-BCCF9F/2020\|EPI_ISL_704405\|20 | ···················· | ···················· | ···················· | ···················· | ······T············· | ···················· | |
| W | 7037 hCoV-19/England/QEUH-BCD5D8/2020\|EPI_ISL_704407\|20 | ···················· | ···················· | ···················· | ···················· | ···················· | ···················· | |
| W | 7038 hCoV-19/England/QEUH-BCC533/2020\|EPI_ISL_704412\|20 | ···················· | ···················· | ···················· | ···················· | ···················· | ···················· | |
| W | 7039 hCoV-19/England/QEUH-BCE23A/2020\|EPI_ISL_704415\|20 | ···················· | ···················· | ···················· | ···················· | ······T············· | ···················· | |
| W | 7040 hCoV-19/England/QEUH-BCC524/2020\|EPI_ISL_704420\|20 | ···················· | ···················· | ···················· | ···················· | ···················· | ···················· | |
| W | 7041 hCoV-19/England/QEUH-BCBB41/2020\|EPI_ISL_704421\|20 | ···················· | ···················· | ···················· | ···················· | ···················· | ···················· | |
| W | 7042 hCoV-19/England/QEUH-BCE382/2020\|EPI_ISL_704423\|20 | ···················· | ···················· | ···················· | ···················· | ···················· | ···················· | |
| W | 7043 hCoV-19/England/QEUH-BCB76B/2020\|EPI_ISL_704424\|20 | ···················· | ···················· | ···················· | ···················· | ···················· | ···················· | |
| W | 7044 hCoV-19/England/QEUH-BCB5CB/2020\|EPI_ISL_704426\|20 | ···················· | ···················· | ···················· | ···················· | ···················· | ···················· | |
| W | 7045 hCoV-19/England/QEUH-BCCCE3/2020\|EPI_ISL_704428\|20 | ···················· | ···················· | ···················· | ···················· | ···················· | ···················· | |
| W | 7046 hCoV-19/England/QEUH-BCC199/2020\|EPI_ISL_704450\|20 | ···················· | ···················· | ···················· | ···················· | ···················· | ···················· | |
| W | 7047 hCoV-19/England/QEUH-BCE540/2020\|EPI_ISL_704451\|20 | ···················· | ···················· | ···················· | ···················· | ···················· | ···················· | |
| W | 7048 hCoV-19/England/QEUH-BCB543/2020\|EPI_ISL_704453\|20 | ···················· | ···················· | ···················· | ···················· | ···················· | ···················· | |
| W | 7049 hCoV-19/England/QEUH-BCE2EF/2020\|EPI_ISL_704455\|20 | ··················T· | ···················· | ···················· | ···················· | ···················· | ···················· | 1 |
| W | 7050 hCoV-19/England/QEUH-BCCF44/2020\|EPI_ISL_704456\|20 | ···················· | ···················· | ···················· | ···················· | ···················· | ···················· | |
| W | 7051 hCoV-19/Scotland/CVR5963/2020\|EPI_ISL_705700\|2020- | ···················· | ···················· | ···················· | ···················· | ···················· | ···················· | |
| W | 7052 hCoV-19/Scotland/CVR5964/2020\|EPI_ISL_705701\|2020- | ···················· | ···················· | ···················· | ···················· | ···················· | ···················· | |
| W | 7053 hCoV-19/Scotland/CVR5965/2020\|EPI_ISL_705702\|2020- | ···················· | ···················· | ···················· | ···················· | ···················· | ···················· | |
| W | 7054 hCoV-19/Scotland/CVR5966/2020\|EPI_ISL_705703\|2020- | ···················· | ···················· | ···················· | ···················· | ···················· | ···················· | |
| W | 7055 hCoV-19/Scotland/CVR5967/2020\|EPI_ISL_705704\|2020- | ···················· | ···················· | ···················· | ···················· | ···················· | ···················· | |
| W | 7056 hCoV-19/Scotland/CVR5968/2020\|EPI_ISL_705705\|2020- | ···················· | ···················· | ···················· | ···················· | ···················· | ···················· | |
| W | 7057 hCoV-19/Scotland/CVR5969/2020\|EPI_ISL_705706\|2020- | ···················· | ···················· | ···················· | ···················· | ···················· | ···················· | |
| W | 7058 hCoV-19/Scotland/CVR5972/2020\|EPI_ISL_705707\|2020- | ···················· | ···················· | ···················· | ···················· | ···················· | ···················· | |
| W | 7059 hCoV-19/Scotland/CVR5975/2020\|EPI_ISL_705708\|2020- | ···················· | ···················· | ···················· | ···················· | ···················· | ···················· | |
| W | 7060 hCoV-19/Scotland/CVR5981/2020\|EPI_ISL_705709\|2020- | ···················· | ···················· | ···················· | ···················· | ···················· | ···················· | |
| W | 7061 hCoV-19/Scotland/CVR5998/2020\|EPI_ISL_705710\|2020- | ···················· | ···················· | ···················· | ···················· | ···················· | ···················· | |
| W | 7062 hCoV-19/Scotland/CVR6006/2020\|EPI_ISL_705711\|2020- | ···················· | ···················· | ···················· | ···················· | ···················· | ···················· | |
| W | 7063 hCoV-19/Scotland/CVR6030/2020\|EPI_ISL_705712\|2020- | ···················· | ···················· | ···················· | ···················· | ···················· | ···················· | |
| W | 7064 hCoV-19/Scotland/CVR6033/2020\|EPI_ISL_705713\|2020- | ···················· | ···················· | ···················· | ···················· | ···················· | ···················· | |
| W | 7065 hCoV-19/Scotland/CVR6038/2020\|EPI_ISL_705714\|2020- | ···················· | ···················· | ···················· | ···················· | ···················· | ···················· | |
| W | 7066 hCoV-19/Scotland/CVR6040/2020\|EPI_ISL_705715\|2020- | ···················· | ···················· | ···················· | ···················· | ···················· | ···················· | |
| W | 7067 hCoV-19/Scotland/CVR6049/2020\|EPI_ISL_705716\|2020- | ···················· | ···················· | ···················· | ···················· | ···················· | ···················· | |
| W | 7068 hCoV-19/Scotland/CVR6051/2020\|EPI_ISL_705717\|2020- | ···················· | ···················· | ···················· | ···················· | ···················· | ···················· | |
| W | 7069 hCoV-19/Scotland/CVR6052/2020\|EPI_ISL_705718\|2020- | ···················· | ···················· | ···················· | ···················· | ···················· | ···················· | |
| W | 7070 hCoV-19/Scotland/CVR6053/2020\|EPI_ISL_705720\|2020- | ···················· | ···················· | ···················· | ···················· | ···················· | ···················· | |
| W | 7071 hCoV-19/Scotland/CVR6055/2020\|EPI_ISL_705721\|2020- | ···················· | ···················· | ···················· | ···················· | ···················· | ···················· | |
| W | 7072 hCoV-19/Scotland/CVR6059/2020\|EPI_ISL_705722\|2020- | ···················· | ···················· | ···················· | ···················· | ···················· | ···················· | |
| W | 7073 hCoV-19/Scotland/CVR6061/2020\|EPI_ISL_705723\|2020- | ···················· | ···················· | ···················· | ···················· | ···················· | ···················· | |
| W | 7074 hCoV-19/Scotland/CVR6063/2020\|EPI_ISL_705724\|2020- | ···················· | ···················· | ···················· | ···················· | ···················· | ···················· | |
| W | 7075 hCoV-19/Scotland/CVR6066/2020\|EPI_ISL_705725\|2020- | ···················· | ···················· | ···················· | ···················· | ···················· | ···················· | |
| W | 7076 hCoV-19/Scotland/CVR6067/2020\|EPI_ISL_705726\|2020- | ···················· | ···················· | ···················· | ···················· | ···················· | ···················· | |
| W | 7077 hCoV-19/Scotland/CVR6068/2020\|EPI_ISL_705727\|2020- | ···················· | ···················· | ···················· | ···················· | ···················· | ···················· | |
| W | 7078 hCoV-19/Scotland/CVR6069/2020\|EPI_ISL_705728\|2020- | ···················· | ···················· | ···················· | ···················· | ···················· | ···················· | |
| W | 7079 hCoV-19/Scotland/CVR6077/2020\|EPI_ISL_705729\|2020- | ···················· | ···················· | ···················· | ···················· | ···················· | ···················· | |
| W | 7080 hCoV-19/Scotland/CVR6092/2020\|EPI_ISL_705730\|2020- | ···················· | ···················· | ···················· | ···················· | ···················· | ···················· | |
| W | 7081 hCoV-19/Scotland/CVR6098/2020\|EPI_ISL_705731\|2020- | ···················· | ···················· | ···················· | ···················· | ···················· | ···················· | |
| W | 7082 hCoV-19/Scotland/CVR6108/2020\|EPI_ISL_705732\|2020- | ···················· | ···················· | ···················· | ···················· | ···················· | ···················· | |
| W | 7083 hCoV-19/Scotland/CVR6116/2020\|EPI_ISL_705733\|2020- | ···················· | ···················· | ···················· | ···················· | ···················· | ···················· | |
| W | 7085 hCoV-19/Scotland/EDB10249/2020\|EPI_ISL_705735\|2020 | ···················· | ···················· | ···················· | ···················· | ···················· | ···················· | |
| W | 7086 hCoV-19/Scotland/EDB10254/2020\|EPI_ISL_705736\|2020 | ···················· | ···················· | ···················· | ···················· | ···················· | ···················· | |
| W | 7087 hCoV-19/Scotland/EDB10261/2020\|EPI_ISL_705737\|2020 | ···················· | ···················· | ···················· | ···················· | ···················· | ···················· | |
| W | 7088 hCoV-19/Scotland/EDB10264/2020\|EPI_ISL_705738\|2020 | ···················· | ···················· | ···················· | ···················· | ···················· | ···················· | |
| W | 7089 hCoV-19/Scotland/EDB10295/2020\|EPI_ISL_705739\|2020 | ···················· | ···················· | ···················· | ···················· | ···················· | ···················· | |
| W | 7090 hCoV-19/Scotland/EDB10296/2020\|EPI_ISL_705740\|2020 | ···················· | ···················· | ···················· | ···················· | ···················· | ···················· | |
| W | 7093 hCoV-19/Scotland/EDB10351/2020\|EPI_ISL_705743\|2020 | ···················· | ···················· | ···················· | ···················· | ···················· | ···················· | |
| W | 7094 hCoV-19/Scotland/EDB10366/2020\|EPI_ISL_705744\|2020 | ···················· | ···················· | ···················· | ···················· | ···················· | ···················· | |
| W | 7095 hCoV-19/Scotland/EDB10372/2020\|EPI_ISL_705745\|2020 | ···················· | ···················· | ···················· | ···················· | ···················· | ···················· | |
| W | 7096 hCoV-19/Scotland/EDB10393/2020\|EPI_ISL_705746\|2020 | ···················· | ···················· | ···················· | ···················· | ···················· | ···················· | |

| | | | | | | | | |
|---|---|---|---|---|---|---|---|---|
| W | 7097 hCoV-19/Scotland/EDB10400/2020\|EPI_ISL_705747\|2020 | ···················· | ···················· | ···················· | ···················· | ···················· | ···················· | |
| W | 7098 hCoV-19/Scotland/EDB10406/2020\|EPI_ISL_705748\|2020 | ···················· | ···················· | ···················· | ···················· | ···················· | ···················· | |
| W | 7099 hCoV-19/Scotland/EDB10409/2020\|EPI_ISL_705749\|2020 | ···················· | ···················· | ···················· | ···················· | ···················· | ···················· | |
| W | 7100 hCoV-19/Scotland/EDB10413/2020\|EPI_ISL_705750\|2020 | ···················· | ···················· | ···················· | ···················· | ···················· | ···················· | |
| W | 7101 hCoV-19/Scotland/EDB10416/2020\|EPI_ISL_705751\|2020 | ···················· | ·A·················· | ···················· | ···················· | ···················· | ···················· | |
| W | 7102 hCoV-19/Scotland/EDB10419/2020\|EPI_ISL_705752\|2020 | ···················· | ···················· | ···················· | ···················· | ···················· | ···················· | |
| W | 7103 hCoV-19/Scotland/EDB10424/2020\|EPI_ISL_705753\|2020 | ···················· | ···················· | ···················· | ···················· | ···················· | ···················· | |
| W | 7104 hCoV-19/Scotland/EDB10427/2020\|EPI_ISL_705754\|2020 | ···················· | ···················· | ···················· | ···················· | ······T············· | ···················· | |
| W | 7105 hCoV-19/Scotland/EDB10429/2020\|EPI_ISL_705755\|2020 | ···················· | ···················· | ···················· | ···················· | ······T············· | ···················· | |
| W | 7106 hCoV-19/Scotland/EDB10432/2020\|EPI_ISL_705756\|2020 | ···················· | ···················· | ···················· | ···················· | ······T············· | ···················· | |
| W | 7108 hCoV-19/Scotland/EDB10437/2020\|EPI_ISL_705758\|2020 | ···················· | ···················· | ···················· | ···················· | ······T············· | ···················· | |
| W | 7109 hCoV-19/Scotland/EDB10439/2020\|EPI_ISL_705759\|2020 | ···················· | ···················· | ···················· | ···················· | ···················· | ···················· | |
| W | 7110 hCoV-19/Scotland/EDB10573/2020\|EPI_ISL_705780\|2020 | ···················· | ···················· | ···················· | ···················· | ···················· | ···················· | |
| W | 7111 hCoV-19/Scotland/EDB10574/2020\|EPI_ISL_705781\|2020 | ···················· | ···················· | ···················· | ···················· | ···················· | ···················· | |
| W | 7112 hCoV-19/Scotland/EDB10578/2020\|EPI_ISL_705782\|2020 | ···················· | ···················· | ···················· | ···················· | ···················· | ···················· | |
| W | 7113 hCoV-19/Scotland/EDB10585/2020\|EPI_ISL_705783\|2020 | ···················· | ···················· | ···················· | ···················· | ···················· | ···················· | |
| W | 7114 hCoV-19/Scotland/EDB10592/2020\|EPI_ISL_705784\|2020 | ···················· | ···················· | ···················· | ···················· | ···················· | ···················· | |
| W | 7115 hCoV-19/Scotland/EDB10597/2020\|EPI_ISL_705785\|2020 | ···················· | ···················· | ···················· | ···················· | ···················· | ···················· | |
| W | 7116 hCoV-19/Scotland/EDB10622/2020\|EPI_ISL_705786\|2020 | ···················· | ···················· | ···················· | ···················· | ···················· | ···················· | |
| W | 7117 hCoV-19/Scotland/EDB10631/2020\|EPI_ISL_705787\|2020 | ···················· | ···················· | ···················· | ···················· | ···················· | ···················· | |
| W | 7118 hCoV-19/Scotland/EDB10639/2020\|EPI_ISL_705789\|2020 | ···················· | ···················· | ···················· | ···················· | ···················· | ···················· | |
| W | 7119 hCoV-19/England/QEUH-BF0A19/2020\|EPI_ISL_709215\|20 | ···················· | ·A·················· | ···················· | ···················· | ···················· | ···A················ | |
| W | 7120 hCoV-19/England/QEUH-BF0BAD/2020\|EPI_ISL_709242\|20 | ···················· | ···················· | ···················· | ···················· | ···················· | ···················· | |
| W | 7121 hCoV-19/England/QEUH-BF115F/2020\|EPI_ISL_709243\|20 | ···················· | ···················· | ···················· | ···················· | ···················· | ···················· | |
| W | 7122 hCoV-19/England/CAMC-BFB47A/2020\|EPI_ISL_709244\|20 | ···················· | ···················· | ···················· | ···················· | ···················· | ···················· | |
| W | 7123 hCoV-19/England/CAMC-BFB4D4/2020\|EPI_ISL_709246\|20 | ···················· | ·A·················· | ···················· | ···················· | ···················· | ···················· | |
| W | 7124 hCoV-19/England/QEUH-BF11AA/2020\|EPI_ISL_709248\|20 | ···················· | ···················· | ···················· | ···················· | ···················· | ···················· | |
| W | 7125 hCoV-19/England/CAMC-BFBB1B/2020\|EPI_ISL_709252\|20 | ···················· | ···················· | ···················· | ···················· | ···················· | ···················· | |
| W | 7126 hCoV-19/England/CAMC-BFBDE8/2020\|EPI_ISL_709256\|20 | ···················· | ···················· | ···················· | ···················· | ······T············· | ···················· | |
| W | 7127 hCoV-19/England/CAMC-BFBE12/2020\|EPI_ISL_709259\|20 | ···················· | ···················· | ···················· | ···················· | ···················· | ···················· | |
| W | 7128 hCoV-19/England/CAMC-BFBE6D/2020\|EPI_ISL_709261\|20 | ···················· | ···················· | ···················· | ···················· | ···················· | ···················· | |
| W | 7129 hCoV-19/England/QEUH-BF0976/2020\|EPI_ISL_709264\|20 | ···················· | ···················· | ···················· | ···················· | ···················· | ···················· | |
| W | 7130 hCoV-19/England/CAMC-BFB89C/2020\|EPI_ISL_709268\|20 | ···················· | ···················· | ···················· | ···················· | ···················· | ···················· | |
| W | 7131 hCoV-19/England/QEUH-BF0C13/2020\|EPI_ISL_709274\|20 | ···················· | ···················· | ···················· | ···················· | ···················· | ···················· | |
| W | 7132 hCoV-19/England/CAMC-BFB8E7/2020\|EPI_ISL_709275\|20 | ···················· | ···················· | ···················· | ···················· | ···················· | ···················· | |
| W | 7133 hCoV-19/England/QEUH-BF117D/2020\|EPI_ISL_709277\|20 | ···················· | ···················· | ···················· | ···················· | ···················· | ···················· | |
| W | 7134 hCoV-19/England/QEUH-BF0B25/2020\|EPI_ISL_709283\|20 | ···················· | ···················· | ···················· | ···················· | ······T············· | ···················· | |
| W | 7135 hCoV-19/England/QEUH-BF09EF/2020\|EPI_ISL_709285\|20 | ···················· | ···················· | ···················· | ···················· | ···················· | ···················· | |
| W | 7136 hCoV-19/England/CAMC-BFB6CF/2020\|EPI_ISL_709286\|20 | ···················· | ···················· | ···················· | ···················· | ···················· | ···················· | |
| W | 7137 hCoV-19/England/CAMC-BFBCFA/2020\|EPI_ISL_709287\|20 | ···················· | ···················· | ···················· | ···················· | ···················· | ···················· | |
| W | 7139 hCoV-19/England/QEUH-BF0C8C/2020\|EPI_ISL_709290\|20 | ···················· | ·A·················· | ···················· | ···················· | ···················· | ···················· | |
| W | 7140 hCoV-19/England/QEUH-BF0B70/2020\|EPI_ISL_709291\|20 | ···················· | ···················· | ···················· | ···················· | ···················· | ···················· | |
| W | 7141 hCoV-19/England/QEUH-BF0C6E/2020\|EPI_ISL_709292\|20 | ···················· | ···················· | ···················· | ···················· | ···················· | ···················· | |
| W | 7142 hCoV-19/England/CAMC-BFBC27/2020\|EPI_ISL_709293\|20 | ···················· | ···················· | ···················· | ···················· | ···················· | ···················· | |
| W | 7143 hCoV-19/England/CAMC-BFB6ED/2020\|EPI_ISL_709294\|20 | ···················· | ···················· | ···················· | ···················· | ···················· | ···················· | |
| W | 7144 hCoV-19/England/ALDP-C06208/2020\|EPI_ISL_709296\|20 | ···················· | ···················· | ···················· | ···················· | ···················· | ···················· | |
| W | 7145 hCoV-19/England/ALDP-C068AC/2020\|EPI_ISL_709298\|20 | ···················· | ···················· | ···················· | ···················· | ···················· | ···················· | |
| W | 7146 hCoV-19/England/ALDP-C06253/2020\|EPI_ISL_709299\|20 | ···················· | ···················· | ···················· | ···················· | ···················· | ···················· | |
| W | 7147 hCoV-19/USA/CO-CDPHE-2004220872/2020\|EPI_ISL_71020 | ···················· | ···················· | ···················· | ···················· | ···················· | ···················· | |
| W | 7151 hCoV-19/USA/CO-CDPHE-2004230207/2020\|EPI_ISL_71020 | ···················· | ···················· | ···················· | ···················· | ···················· | ···················· | |
| W | 7152 hCoV-19/USA/CO-CDPHE-2009151365/2020\|EPI_ISL_71022 | ···················· | ···················· | ···················· | ···················· | ···················· | ···················· | |
| W | 7153 hCoV-19/USA/CO-CDPHE-2009151379/2020\|EPI_ISL_71022 | ···················· | ···················· | ···················· | ···················· | ···················· | ···················· | |
| W | 7154 hCoV-19/USA/CO-CDPHE-2009151427/2020\|EPI_ISL_71023 | ···················· | ···················· | ···················· | ···················· | ···················· | ···················· | |
| W | 7155 hCoV-19/USA/CO-CDPHE-2009163897/2020\|EPI_ISL_71023 | ···················· | ·A·················· | ···················· | ···················· | ···················· | ···················· | |
| W | 7156 hCoV-19/USA/CO-CDPHE-2009163949/2020\|EPI_ISL_71023 | ···················· | ·A·················· | ···················· | ···················· | ···················· | ···················· | |
| W | 7157 hCoV-19/USA/CO-CDPHE-2009163956/2020\|EPI_ISL_71023 | ···················· | ·A·················· | ···················· | ···················· | ···················· | ···················· | |
| W | 7158 hCoV-19/USA/CO-CDPHE-2009163959/2020\|EPI_ISL_71023 | ···················· | ···················· | ···················· | ···················· | ···················· | ···················· | |
| W | 7159 hCoV-19/USA/CO-CDPHE-2009164005/2020\|EPI_ISL_71023 | ···················· | ···················· | ···················· | ···················· | ···················· | ···················· | |
| W | 7160 hCoV-19/USA/CO-CDPHE-2009164052/2020\|EPI_ISL_71023 | ···················· | ·A·················· | ···················· | ···················· | ···················· | ···················· | |
| W | 7161 hCoV-19/USA/CO-CDPHE-2010200348/2020\|EPI_ISL_71023 | ···················· | ···················· | ···················· | ···················· | ···················· | ···················· | |
| W | 7162 hCoV-19/USA/CO-CDPHE-2010200390/2020\|EPI_ISL_71023 | ···················· | ···················· | ···················· | ···················· | ···················· | ···················· | |
| W | 7163 hCoV-19/USA/CO-CDPHE-2010200579/2020\|EPI_ISL_71023 | ···················· | ···················· | ···················· | ···················· | ···················· | ···················· | |
| W | 7164 hCoV-19/USA/CO-CDPHE-2010280225/2020\|EPI_ISL_71024 | ···················· | ···················· | ···················· | ···················· | NNN················· | ···················· | 1 |
| W | 7166 hCoV-19/USA/CO-CDPHE-2011020172/2020\|EPI_ISL_71024 | ···················· | ···················· | ···················· | ···················· | ···················· | ···················· | |
| W | 7167 hCoV-19/USA/CO-CDPHE-2011020202/2020\|EPI_ISL_71024 | ···················· | ···················· | ···················· | ···················· | ···················· | ···················· | |
| W | 7168 hCoV-19/USA/CO-CDPHE-2011020245/2020\|EPI_ISL_71024 | ···················· | ···················· | ···················· | ···················· | ···················· | ···················· | |
| W | 7169 hCoV-19/USA/AZ-TG395909/2020\|EPI_ISL_694753\|2020-0 | ···················· | ···················· | ···················· | ···················· | ···················· | ···················· | |
| W | 7170 hCoV-19/USA/AZ-TG395911/2020\|EPI_ISL_694755\|2020-0 | ···················· | ···················· | ···················· | ···················· | ···················· | ···················· | |
| W | 7171 hCoV-19/USA/AZ-TG395915/2020\|EPI_ISL_694758\|2020-0 | ···················· | ···················· | ···················· | ···················· | ···················· | ···················· | |
| W | 7172 hCoV-19/USA/AZ-TG395920/2020\|EPI_ISL_694761\|2020-0 | ···················· | ···················· | ···················· | ···················· | ···················· | ···················· | |
| W | 7173 hCoV-19/USA/AZ-TG395923/2020\|EPI_ISL_694763\|2020-0 | ···················· | ···················· | ···················· | ···················· | ···················· | ···················· | |
| W | 7174 hCoV-19/USA/AZ-TG395928/2020\|EPI_ISL_694764\|2020-0 | ···················· | ···················· | ···················· | ···················· | ···················· | ···················· | |
| W | 7175 hCoV-19/USA/AZ-TG395929/2020\|EPI_ISL_694765\|2020-0 | ···················· | ···················· | ···················· | ···················· | ···················· | ···················· | |
| W | 7176 hCoV-19/USA/AZ-TG395933/2020\|EPI_ISL_694769\|2020-0 | ···················· | ···················· | ···················· | ···················· | ···················· | ···················· | |
| W | 7177 hCoV-19/USA/AZ-TG395935/2020\|EPI_ISL_694771\|2020-0 | ···················· | ·············A······ | ···················· | ···················· | ···················· | ···················· | |
| W | 7178 hCoV-19/USA/AZ-TG395937/2020\|EPI_ISL_694772\|2020-0 | ···················· | ·A·················· | ···················· | ···················· | ···················· | ···················· | |
| W | 7179 hCoV-19/USA/AZ-TG395938/2020\|EPI_ISL_694773\|2020-0 | ···················· | ···················· | ···················· | ···················· | ···················· | ···················· | |
| W | 7180 hCoV-19/USA/AZ-TG395940/2020\|EPI_ISL_694775\|2020-0 | ···················· | ···················· | ···················· | ···················· | ···················· | ···················· | |
| W | 7181 hCoV-19/USA/AZ-TG395948/2020\|EPI_ISL_694781\|2020-0 | ···················· | ···················· | ···················· | ···················· | ···················· | ···················· | |
| W | 7182 hCoV-19/USA/AZ-TG395951/2020\|EPI_ISL_694782\|2020-0 | ···················· | ···················· | ···················· | ···················· | ···················· | ···················· | |

| | | | | | | | | |
|---|---|---|---|---|---|---|---|---|
| W | 7183 | hCoV-19/USA/AZ-TG395958/2020\|EPI_ISL_694784\|2020-0 | ···················· | ···················· | ···················· | ···················· | ···················· | ···················· |
| W | 7184 | hCoV-19/USA/AZ-TG395960/2020\|EPI_ISL_694786\|2020-0 | ···················· | ·A·················· | ···················· | ···················· | ···················· | ···················· |
| W | 7185 | hCoV-19/USA/AZ-TG395963/2020\|EPI_ISL_694788\|2020-0 | ···················· | ·A·················· | ···················· | ···················· | ···················· | ···················· |
| W | 7186 | hCoV-19/USA/AZ-TG395971/2020\|EPI_ISL_694796\|2020-0 | ···················· | ···················· | ···················· | ···················· | ···················· | ···················· |
| W | 7187 | hCoV-19/USA/AZ-TG396023/2020\|EPI_ISL_694797\|2020-0 | ···················· | ···················· | ···················· | ···················· | ···················· | ···················· |
| W | 7189 | hCoV-19/United Arab Emirates/0110/2020\|EPI_ISL_698 | ···················· | ···················· | ···················· | ···················· | ···················· | ···················· |
| W | 7190 | hCoV-19/United Arab Emirates/0111/2020\|EPI_ISL_698 | ···················· | ···················· | ···················· | ···················· | ···················· | ···················· |
| W | 7191 | hCoV-19/United Arab Emirates/0112/2020\|EPI_ISL_698 | ···················· | ···················· | ···················· | ···················· | ···················· | ···················· |
| W | 7192 | hCoV-19/United Arab Emirates/0113/2020\|EPI_ISL_698 | ···················· | ···················· | ···················· | ···················· | ···················· | ···················· |
| W | 7193 | hCoV-19/United Arab Emirates/0114/2020\|EPI_ISL_698 | ···················· | ···················· | ···················· | ···················· | ···················· | ···················· |
| W | 7194 | hCoV-19/United Arab Emirates/0115/2020\|EPI_ISL_698 | ···················· | ···················· | ···················· | ···················· | ···················· | ···················· |
| W | 7195 | hCoV-19/United Arab Emirates/0116/2020\|EPI_ISL_698 | ···················· | ···················· | ···················· | ···················· | ···················· | ···················· |
| W | 7196 | hCoV-19/United Arab Emirates/0117/2020\|EPI_ISL_698 | ···················· | ···················· | ···················· | ···················· | ···················· | ···················· |
| W | 7197 | hCoV-19/United Arab Emirates/0118/2020\|EPI_ISL_698 | ···················· | ···················· | ···················· | ···················· | ···················· | ···················· |
| W | 7198 | hCoV-19/United Arab Emirates/0119/2020\|EPI_ISL_698 | ···················· | ···················· | ···················· | ···················· | ···················· | ···················· |
| W | 7199 | hCoV-19/United Arab Emirates/0120/2020\|EPI_ISL_698 | ···················· | ···················· | ···················· | ···················· | ···················· | ···················· |
| W | 7200 | hCoV-19/United Arab Emirates/0121/2020\|EPI_ISL_698 | ···················· | ···················· | ···················· | ···················· | ···················· | ···················· |
| W | 7201 | hCoV-19/United Arab Emirates/0122/2020\|EPI_ISL_698 | ···················· | ···················· | ···················· | ···················· | ···················· | ···················· |
| W | 7202 | hCoV-19/United Arab Emirates/0123/2020\|EPI_ISL_698 | ···················· | ···················· | ···················· | ···················· | ···················· | ···················· |
| W | 7203 | hCoV-19/United Arab Emirates/0124/2020\|EPI_ISL_698 | ···················· | ···················· | ···················· | ···················· | ···················· | ···················· |
| W | 7204 | hCoV-19/United Arab Emirates/0173/2020\|EPI_ISL_698 | ···················· | ···················· | ···················· | ···················· | ···················· | ···················· |
| W | 7205 | hCoV-19/United Arab Emirates/0174/2020\|EPI_ISL_698 | ···················· | ···················· | ···················· | ···················· | ···················· | ···················· |
| W | 7206 | hCoV-19/United Arab Emirates/0183/2020\|EPI_ISL_698 | ···················· | ···················· | ···················· | ···················· | ···················· | ···················· |
| W | 7207 | hCoV-19/United Arab Emirates/0184/2020\|EPI_ISL_698 | ···················· | ···················· | ···················· | ···················· | ···················· | ···················· |
| W | 7208 | hCoV-19/United Arab Emirates/0185/2020\|EPI_ISL_698 | ···················· | ···················· | ···················· | ···················· | ···················· | ···················· |
| W | 7209 | hCoV-19/United Arab Emirates/0186/2020\|EPI_ISL_698 | ···················· | ···················· | ···················· | ···················· | ···················· | ···················· |
| W | 7210 | hCoV-19/United Arab Emirates/0187/2020\|EPI_ISL_698 | ···················· | ···················· | ···················· | ···················· | ···················· | ···················· |
| W | 7211 | hCoV-19/United Arab Emirates/0189/2020\|EPI_ISL_698 | ···················· | ···················· | ···················· | ···················· | ···················· | ···················· |
| W | 7212 | hCoV-19/United Arab Emirates/0190/2020\|EPI_ISL_698 | ···················· | ···················· | ···················· | ···················· | ···················· | ···················· |
| W | 7213 | hCoV-19/United Arab Emirates/0191/2020\|EPI_ISL_698 | ···················· | ···················· | ···················· | ···················· | ···················· | ···················· |
| W | 7214 | hCoV-19/United Arab Emirates/0192/2020\|EPI_ISL_698 | ···················· | ···················· | ···················· | ···················· | ···················· | ···················· |
| W | 7215 | hCoV-19/United Arab Emirates/0193/2020\|EPI_ISL_698 | ···················· | ·A·················· | ···················· | ···················· | ···················· | ···················· |
| W | 7216 | hCoV-19/United Arab Emirates/0194/2020\|EPI_ISL_698 | ···················· | ···················· | ···················· | ···················· | ···················· | ···················· |
| W | 7217 | hCoV-19/United Arab Emirates/0195/2020\|EPI_ISL_698 | ···················· | ···················· | ···················· | ···················· | ···················· | ···················· |
| W | 7218 | hCoV-19/United Arab Emirates/0196/2020\|EPI_ISL_698 | ···················· | ···················· | ···················· | ···················· | ···················· | ···················· |
| W | 7219 | hCoV-19/United Arab Emirates/0197/2020\|EPI_ISL_698 | ···················· | ···················· | ···················· | ···················· | ···················· | ···················· |
| W | 7220 | hCoV-19/United Arab Emirates/0198/2020\|EPI_ISL_698 | ···················· | ···················· | ···················· | ···················· | ···················· | ···················· |
| W | 7221 | hCoV-19/United Arab Emirates/0199/2020\|EPI_ISL_698 | ···················· | ···················· | ···················· | ···················· | ···················· | ···················· |
| W | 7222 | hCoV-19/United Arab Emirates/0200/2020\|EPI_ISL_698 | ···················· | ···················· | ···················· | ···················· | ···················· | ···················· |
| W | 7223 | hCoV-19/United Arab Emirates/0201/2020\|EPI_ISL_698 | ···················· | ···················· | ···················· | ···················· | ···················· | ···················· |
| W | 7224 | hCoV-19/United Arab Emirates/0202/2020\|EPI_ISL_698 | ···················· | ···················· | ···················· | ···················· | ···················· | ···················· |
| W | 7225 | hCoV-19/United Arab Emirates/0203/2020\|EPI_ISL_698 | ···················· | ···················· | ···················· | ···················· | ···················· | ···················· |
| W | 7226 | hCoV-19/United Arab Emirates/0204/2020\|EPI_ISL_698 | ···················· | ···················· | ···················· | ···················· | ···················· | ···················· |
| W | 7227 | hCoV-19/United Arab Emirates/0205/2020\|EPI_ISL_698 | ···················· | ···················· | ···················· | ···················· | ···················· | ···················· |
| W | 7228 | hCoV-19/United Arab Emirates/0206/2020\|EPI_ISL_698 | ···················· | ···················· | ···················· | ···················· | ···················· | ···················· |
| W | 7229 | hCoV-19/United Arab Emirates/0207/2020\|EPI_ISL_698 | ···················· | ···················· | ···················· | ···················· | ···················· | ···················· |
| W | 7230 | hCoV-19/United Arab Emirates/0208/2020\|EPI_ISL_698 | ···················· | ···················· | ···················· | ···················· | ···················· | ···················· |
| W | 7231 | hCoV-19/United Arab Emirates/0209/2020\|EPI_ISL_698 | ···················· | ···················· | ···················· | ···················· | ···················· | ···················· |
| W | 7232 | hCoV-19/United Arab Emirates/0210/2020\|EPI_ISL_698 | ···················· | ···················· | ···················· | ···················· | ···················· | ···················· |
| W | 7233 | hCoV-19/United Arab Emirates/0211/2020\|EPI_ISL_698 | ···················· | ···················· | ···················· | ···················· | ···················· | ···················· |
| W | 7234 | hCoV-19/United Arab Emirates/0212/2020\|EPI_ISL_698 | ···················· | ···················· | ···················· | ···················· | ···················· | ···················· |
| W | 7235 | hCoV-19/United Arab Emirates/0213/2020\|EPI_ISL_698 | ···················· | ···················· | ···················· | ···················· | ···················· | ···················· |
| W | 7236 | hCoV-19/India/MH-ACTREC-034/2020\|EPI_ISL_699690\|20 | ···················· | ···················· | ···················· | ···················· | ···················· | ···················· |
| W | 7237 | hCoV-19/England/ALDP-BB52CD/2020\|EPI_ISL_703100\|20 | ···················· | ···················· | ···················· | ···················· | ···················· | ···················· |
| W | 7238 | hCoV-19/England/ALDP-BB2A31/2020\|EPI_ISL_703101\|20 | ···················· | ···················· | ···················· | ···················· | ···················· | ···················· |
| W | 7239 | hCoV-19/England/ALDP-BB2B10/2020\|EPI_ISL_703103\|20 | ···················· | ···················· | ···················· | ···················· | ···················· | ···················· |
| W | 7240 | hCoV-19/England/ALDP-BB555B/2020\|EPI_ISL_703106\|20 | ···················· | ···················· | ···················· | ···················· | ···················· | ···················· |
| W | 7241 | hCoV-19/England/ALDP-BB3292/2020\|EPI_ISL_703108\|20 | ···················· | ···················· | ···················· | ···················· | ···················· | ···················· |
| W | 7242 | hCoV-19/England/ALDP-BB34BA/2020\|EPI_ISL_703109\|20 | ···················· | ···················· | ···················· | ···················· | ······T············· | ···················· |
| W | 7243 | hCoV-19/England/ALDP-BB28FB/2020\|EPI_ISL_703111\|20 | ···················· | ···················· | ···················· | ···················· | ···················· | ···················· |
| W | 7244 | hCoV-19/England/ALDP-BB52DC/2020\|EPI_ISL_703112\|20 | ···················· | ···················· | ···················· | ···················· | ···················· | ···················· |
| W | 7245 | hCoV-19/England/ALDP-BB398E/2020\|EPI_ISL_703114\|20 | ···················· | ···················· | ···················· | ···················· | ···················· | ···················· |
| W | 7246 | hCoV-19/England/ALDP-BB2BE3/2020\|EPI_ISL_703116\|20 | ···················· | ···················· | ···················· | ···················· | ···················· | ···················· |
| W | 7247 | hCoV-19/England/ALDP-BB3A03/2020\|EPI_ISL_703117\|20 | ···················· | ···················· | ···················· | ···················· | ···················· | ···················· |
| W | 7248 | hCoV-19/England/ALDP-BB8DE7/2020\|EPI_ISL_703131\|20 | ···················· | ···················· | ···················· | ···················· | ···················· | ···················· |
| W | 7249 | hCoV-19/England/ALDP-BB3502/2020\|EPI_ISL_703134\|20 | ···················· | ···················· | ···················· | ···················· | ···················· | ···················· |
| W | 7250 | hCoV-19/England/ALDP-BB9B19/2020\|EPI_ISL_703135\|20 | ···················· | ···················· | ···················· | ···················· | ···················· | ···················· |
| W | 7251 | hCoV-19/England/ALDP-BB9AB2/2020\|EPI_ISL_703142\|20 | ···················· | ···················· | ···················· | ···················· | ···················· | ···················· |
| W | 7252 | hCoV-19/England/ALDP-BB8E02/2020\|EPI_ISL_703143\|20 | ···················· | ·········T·········· | ···················· | ···················· | ···················· | ···················· |
| W | 7253 | hCoV-19/England/ALDP-BB2A22/2020\|EPI_ISL_703145\|20 | ···················· | ···················· | ···················· | ···················· | ···················· | ···················· |
| W | 7254 | hCoV-19/England/ALDP-BB9D7D/2020\|EPI_ISL_703146\|20 | ···················· | ···················· | ···················· | ···················· | ···················· | ···················· |
| W | 7255 | hCoV-19/England/ALDP-BB9496/2020\|EPI_ISL_703149\|20 | ···················· | ···················· | ···················· | ···················· | ···················· | ···················· |
| W | 7256 | hCoV-19/England/ALDP-BB554C/2020\|EPI_ISL_703153\|20 | ···················· | ···················· | ···················· | ···················· | ···················· | ···················· |
| W | 7257 | hCoV-19/England/ALDP-BBACCA/2020\|EPI_ISL_703156\|20 | ···················· | ···················· | ···················· | ···················· | ···················· | ···················· |
| W | 7258 | hCoV-19/England/ALDP-BB3414/2020\|EPI_ISL_703157\|20 | ···················· | ···················· | ···················· | ···················· | ······T············· | ···················· |
| W | 7259 | hCoV-19/England/ALDP-BB8BDE/2020\|EPI_ISL_703159\|20 | ···················· | ···················· | ···················· | ···················· | ···················· | ···················· |
| W | 7260 | hCoV-19/England/ALDP-BB945A/2020\|EPI_ISL_703160\|20 | ···················· | ···················· | ···················· | ···················· | ···················· | ···················· |
| W | 7261 | hCoV-19/England/ALDP-BB2B98/2020\|EPI_ISL_703161\|20 | ···················· | ···················· | ···················· | ···················· | ···················· | ···················· |
| W | 7262 | hCoV-19/England/ALDP-BB5764/2020\|EPI_ISL_703163\|20 | ···················· | ···················· | ···················· | ···················· | ···················· | ···················· |
| W | 7263 | hCoV-19/England/ALDP-BB2907/2020\|EPI_ISL_703166\|20 | ···················· | ···················· | ···················· | ···················· | ······T············· | ···················· |

| | | | | | | | | |
|---|---|---|---|---|---|---|---|---|
| W | 7264 | hCoV-19/England/ALDP-BB5DEA/2020\|EPI_ISL_703168\|20 | ···················· | ···················· | ····················· | ···················· | ···················· | ···················· |
| W | 7265 | hCoV-19/England/ALDP-BB3168/2020\|EPI_ISL_703171\|20 | ···················· | ···················· | ····················· | ···················· | ···················· | ···················· |
| W | 7266 | hCoV-19/England/ALDP-BB56FE/2020\|EPI_ISL_703173\|20 | ···················· | ···················· | ····················· | ···················· | ···················· | ···················· |
| W | 7267 | hCoV-19/England/ALDP-BB370C/2020\|EPI_ISL_703175\|20 | ···················· | ···················· | ····················· | ···················· | ···················· | ···················· |
| W | 7268 | hCoV-19/England/ALDP-BB94A5/2020\|EPI_ISL_703183\|20 | ···················· | ···················· | ····················· | ···················· | ···················· | ···················· |
| W | 7269 | hCoV-19/England/ALDP-BB39F7/2020\|EPI_ISL_703185\|20 | ···················· | ···················· | ····················· | ···················· | ···················· | ···················· |
| W | 7270 | hCoV-19/England/ALDP-BB9D40/2020\|EPI_ISL_703186\|20 | ···················· | ···················· | ····················· | ···················· | ···················· | ···················· |
| W | 7271 | hCoV-19/England/ALDP-BBACAC/2020\|EPI_ISL_703188\|20 | ···················· | ···················· | ····················· | ···················· | ···················· | ···················· |
| W | 7272 | hCoV-19/England/ALDP-BB2873/2020\|EPI_ISL_703189\|20 | ···················· | ···················· | ····················· | ···················· | ······T············· | ···················· |
| W | 7273 | hCoV-19/England/ALDP-BB33DB/2020\|EPI_ISL_703191\|20 | ···················· | ···················· | ····················· | ···················· | ···················· | ···················· |
| W | 7274 | hCoV-19/England/ALDP-BB2819/2020\|EPI_ISL_703193\|20 | ···················· | ···················· | ····················· | ···················· | ···················· | ···················· |
| W | 7275 | hCoV-19/England/ALDP-BB2DDE/2020\|EPI_ISL_703194\|20 | ···················· | ···················· | ····················· | ···················· | ······T············· | ···················· |
| W | 7276 | hCoV-19/England/ALDP-BB2BA7/2020\|EPI_ISL_703199\|20 | ···················· | ···················· | ····················· | ···················· | ···················· | ···················· |
| W | 7277 | hCoV-19/England/QEUH-BCBA53/2020\|EPI_ISL_704431\|20 | ···················· | ···················· | ····················· | ···················· | ···················· | ···················· |
| W | 7278 | hCoV-19/England/QEUH-BCE2FE/2020\|EPI_ISL_704434\|20 | ···················· | ···················· | ····················· | ···················· | ···················· | ···················· |
| W | 7279 | hCoV-19/England/QEUH-BCDA42/2020\|EPI_ISL_704440\|20 | ···················· | ···················· | ····················· | ···················· | ···················· | ···················· |
| W | 7280 | hCoV-19/England/QEUH-BCB525/2020\|EPI_ISL_704442\|20 | ···················· | ···················· | ····················· | ···················· | ···················· | ···················· |
| W | 7281 | hCoV-19/England/QEUH-BCE892/2020\|EPI_ISL_704444\|20 | ···················· | ···················· | ····················· | ···················· | ···················· | ···················· |
| W | 7282 | hCoV-19/England/QEUH-BCCFDB/2020\|EPI_ISL_704445\|20 | ···················· | ·A·················· | ····················· | ···················· | ···················· | ···················· |
| W | 7283 | hCoV-19/England/QEUH-BCD6C6/2020\|EPI_ISL_704447\|20 | ···················· | ···················· | ····················· | ···················· | ···················· | ···················· |
| W | 7284 | hCoV-19/England/QEUH-BCBB6F/2020\|EPI_ISL_704448\|20 | ···················· | ···················· | ····················· | ···················· | ···················· | ···················· |
| W | 7285 | hCoV-19/England/QEUH-BCBDB4/2020\|EPI_ISL_704458\|20 | ···················· | ···················· | ····················· | ···················· | ···················· | ···················· |
| W | 7286 | hCoV-19/England/QEUH-BCDD2B/2020\|EPI_ISL_704461\|20 | ···················· | ···················· | ····················· | ···················· | ···················· | ···················· |
| W | 7287 | hCoV-19/England/QEUH-BCD7D2/2020\|EPI_ISL_704463\|20 | ···················· | ···················· | ····················· | ···················· | ······T············· | ···················· |
| W | 7288 | hCoV-19/England/QEUH-BCE1C4/2020\|EPI_ISL_704464\|20 | ···················· | ···················· | ····················· | ···················· | ···················· | ···················· |
| W | 7289 | hCoV-19/England/QEUH-BCD7C3/2020\|EPI_ISL_704466\|20 | ···················· | ···················· | ····················· | ···················· | ···················· | ···················· |
| W | 7290 | hCoV-19/England/QEUH-BCD67B/2020\|EPI_ISL_704468\|20 | ···················· | ···················· | ····················· | ···················· | ···················· | ···················· |
| W | 7291 | hCoV-19/England/QEUH-BCD981/2020\|EPI_ISL_704469\|20 | ···················· | ···················· | ····················· | ···················· | ···················· | ···················· |
| W | 7292 | hCoV-19/England/QEUH-BCC542/2020\|EPI_ISL_704471\|20 | ···················· | ···················· | ····················· | ···················· | ···················· | ···················· |
| W | 7293 | hCoV-19/England/QEUH-BCB6E6/2020\|EPI_ISL_704474\|20 | ···················· | ···················· | ····················· | ···················· | ···················· | ···················· |
| W | 7294 | hCoV-19/England/QEUH-BCB570/2020\|EPI_ISL_704476\|20 | ···················· | ···················· | ····················· | ···················· | ···················· | ···················· |
| W | 7295 | hCoV-19/England/QEUH-BCD66C/2020\|EPI_ISL_704477\|20 | ···················· | ···················· | ····················· | ···················· | ···················· | ···················· |
| W | 7296 | hCoV-19/England/QEUH-BCD9EB/2020\|EPI_ISL_704479\|20 | ···················· | ···················· | ····················· | ···················· | ···················· | ···················· |
| W | 7297 | hCoV-19/England/QEUH-BCE522/2020\|EPI_ISL_704480\|20 | ···················· | ···················· | ····················· | ···················· | ···················· | ···················· |
| W | 7298 | hCoV-19/England/QEUH-BCCDB3/2020\|EPI_ISL_704482\|20 | ···················· | ···················· | ····················· | ···················· | ···················· | ···················· |
| W | 7299 | hCoV-19/England/QEUH-BCC17B/2020\|EPI_ISL_704483\|20 | ···················· | ···················· | ····················· | ···················· | ···················· | ···················· |
| W | 7300 | hCoV-19/England/QEUH-BCCE83/2020\|EPI_ISL_704485\|20 | ···················· | ···················· | ····················· | ···················· | ···················· | ···················· |
| W | 7301 | hCoV-19/England/QEUH-BCD71E/2020\|EPI_ISL_704487\|20 | ···················· | ···················· | ····················· | ···················· | ···················· | ···················· |
| W | 7302 | hCoV-19/England/QEUH-BCD417/2020\|EPI_ISL_704488\|20 | ···················· | ···················· | ····················· | ···················· | ···················· | ···················· |
| W | 7303 | hCoV-19/England/QEUH-BCD9AF/2020\|EPI_ISL_704491\|20 | ···················· | ···················· | ····················· | ···················· | ···················· | ···················· |
| W | 7304 | hCoV-19/England/QEUH-BCE917/2020\|EPI_ISL_704493\|20 | ···················· | ···················· | ····················· | ···················· | ···················· | ···················· |
| W | 7305 | hCoV-19/England/QEUH-BCBDF0/2020\|EPI_ISL_704495\|20 | ···················· | ···················· | ····················· | ···················· | ···················· | ···················· |
| W | 7306 | hCoV-19/England/QEUH-BCE8CF/2020\|EPI_ISL_704496\|20 | ···················· | ···················· | ····················· | ···················· | ···················· | ···················· |
| W | 7307 | hCoV-19/England/QEUH-BCE953/2020\|EPI_ISL_704498\|20 | ···················· | ···················· | ····················· | ···················· | ···················· | ···················· |
| W | 7308 | hCoV-19/Scotland/EDB10442/2020\|EPI_ISL_705760\|2020 | ···················· | ···················· | ····················· | ···················· | ······T············· | ···················· |
| W | 7309 | hCoV-19/Scotland/EDB10445/2020\|EPI_ISL_705761\|2020 | ···················· | ···················· | ····················· | ···················· | ···················· | ···················· |
| W | 7310 | hCoV-19/Scotland/EDB10447/2020\|EPI_ISL_705762\|2020 | ···················· | ···················· | ····················· | ···················· | ···················· | ···················· |
| W | 7311 | hCoV-19/Scotland/EDB10448/2020\|EPI_ISL_705763\|2020 | ···················· | ···················· | ····················· | ···················· | ···················· | ···················· |
| W | 7312 | hCoV-19/Scotland/EDB10451/2020\|EPI_ISL_705764\|2020 | ···················· | ···················· | ····················· | ···················· | ···················· | ···················· |
| W | 7313 | hCoV-19/Scotland/EDB10455/2020\|EPI_ISL_705765\|2020 | ···················· | ···················· | ····················· | ···················· | ······T············· | ···················· |
| W | 7314 | hCoV-19/Scotland/EDB10459/2020\|EPI_ISL_705766\|2020 | ···················· | ···················· | ····················· | ···················· | ···················· | ···················· |
| W | 7315 | hCoV-19/Scotland/EDB10463/2020\|EPI_ISL_705767\|2020 | ···················· | ···················· | ····················· | ···················· | ···················· | ···················· |
| W | 7316 | hCoV-19/Scotland/EDB10471/2020\|EPI_ISL_705768\|2020 | ···················· | ···················· | ····················· | ···················· | ···················· | ···················· |
| W | 7317 | hCoV-19/Scotland/EDB10472/2020\|EPI_ISL_705769\|2020 | ···················· | ···················· | ····················· | ···················· | ···················· | ···A················ |
| W | 7318 | hCoV-19/Scotland/EDB10475/2020\|EPI_ISL_705770\|2020 | ···················· | ···················· | ····················· | ···················· | ···················· | ···················· |
| W | 7319 | hCoV-19/Scotland/EDB10479/2020\|EPI_ISL_705771\|2020 | ···················· | ···················· | ····················· | ···················· | ···················· | ···················· |
| W | 7320 | hCoV-19/Scotland/EDB10480/2020\|EPI_ISL_705772\|2020 | ···················· | ···················· | ····················· | ···················· | ···················· | ···················· |
| W | 7321 | hCoV-19/Scotland/EDB10483/2020\|EPI_ISL_705773\|2020 | ···················· | ···················· | ····················· | ···················· | ···················· | ···················· |
| W | 7323 | hCoV-19/Scotland/EDB10517/2020\|EPI_ISL_705775\|2020 | ···················· | ···················· | ····················· | ···················· | ···················· | ···················· |
| W | 7325 | hCoV-19/Scotland/EDB10520/2020\|EPI_ISL_705777\|2020 | ···················· | ···················· | ····················· | ···················· | ···················· | ···················· |
| W | 7326 | hCoV-19/Scotland/EDB10553/2020\|EPI_ISL_705778\|2020 | ···················· | ···················· | ····················· | ···················· | ···················· | ···················· |
| W | 7327 | hCoV-19/Scotland/EDB10557/2020\|EPI_ISL_705779\|2020 | ···················· | ···················· | ····················· | ···················· | ···················· | ···················· |
| W | 7328 | hCoV-19/Scotland/EDB10633/2020\|EPI_ISL_705788\|2020 | ···················· | ···················· | ····················· | ···················· | ···················· | ···················· |
| W | 7329 | hCoV-19/Scotland/EDB10643/2020\|EPI_ISL_705790\|2020 | ···················· | ···················· | ····················· | ···················· | ···················· | ···················· |
| W | 7330 | hCoV-19/Scotland/EDB10645/2020\|EPI_ISL_705791\|2020 | ···················· | ···················· | ····················· | ···················· | ···················· | ···················· |
| W | 7331 | hCoV-19/Scotland/EDB10658/2020\|EPI_ISL_705792\|2020 | ···················· | ···················· | ····················· | ···················· | ···················· | ···················· |
| W | 7332 | hCoV-19/Scotland/EDB10674/2020\|EPI_ISL_705793\|2020 | ···················· | ···················· | ····················· | ···················· | ···················· | ···················· |
| W | 7333 | hCoV-19/Scotland/EDB10724/2020\|EPI_ISL_705794\|2020 | ···················· | ···················· | ····················· | ···················· | ···················· | ···················· |
| W | 7334 | hCoV-19/Scotland/EDB1269/2020\|EPI_ISL_705795\|2020- | ···················· | ···················· | ····················· | ···················· | ···················· | ···················· |
| W | 7336 | hCoV-19/Scotland/EDB741/2020\|EPI_ISL_705797\|2020-0 | ···················· | ·A·················· | ····················· | ···················· | ···················· | ···················· |
| W | 7337 | hCoV-19/Scotland/EDB9053/2020\|EPI_ISL_705798\|2020- | ···················· | ···················· | ····················· | ···················· | ···················· | ···················· |
| W | 7338 | hCoV-19/Scotland/EDB9268/2020\|EPI_ISL_705799\|2020- | ···················· | ···················· | ····················· | ···················· | ······T············· | ···················· |
| W | 7339 | hCoV-19/Germany/SL-SU-10452452/2020\|EPI_ISL_708000 | ···················· | ···················· | ····················· | ···················· | ······T············· | ···················· |
| W | 7340 | hCoV-19/Germany/SL-SU-20180793/2020\|EPI_ISL_708003 | ···················· | ···················· | ····················· | ···················· | ···················· | ···················· |
| W | 7342 | hCoV-19/Germany/SL-SU-20241112/2020\|EPI_ISL_708014 | ···················· | ···················· | ····················· | ···················· | ···················· | ···················· |
| W | 7343 | hCoV-19/Germany/SL-SU-20242379/2020\|EPI_ISL_708015 | ···················· | ···················· | ····················· | ···················· | ···················· | ···················· |
| W | 7344 | hCoV-19/Norway/3576/2020\|EPI_ISL_708026\|2020-08-06 | ···················· | ·A·················· | ····················· | ···················· | ···················· | ···A················ |
| W | 7345 | hCoV-19/Norway/3597/2020\|EPI_ISL_708027\|2020-07-31 | ···················· | ·A·················· | ····················· | ···················· | ···················· | ···A················ |
| W | 7346 | hCoV-19/Norway/4288/2020\|EPI_ISL_708028\|2020-10-13 | ···················· | ···················· | ····················· | ···················· | ···················· | ···················· |
| W | 7347 | hCoV-19/Norway/4474/2020\|EPI_ISL_708029\|2020-10-18 | ···················· | ···················· | ····················· | ···················· | ···················· | ···················· |

| | | | | | | | | |
|---|---|---|---|---|---|---|---|---|
| W | 7348 hCoV-19/England/ALDP-C061FC/2020\|EPI_ISL_709300\|20 | ···················· | ···················· | ···················· | ···················· | ···················· | ···················· | |
| W | 7349 hCoV-19/England/QEUH-C163D5/2020\|EPI_ISL_709301\|20 | ···················· | ···················· | ···················· | ···················· | ···················· | ···················· | |
| W | 7350 hCoV-19/England/ALDP-C062EA/2020\|EPI_ISL_709302\|20 | ···················· | ···················· | ···················· | ···················· | ···················· | ···················· | |
| W | 7351 hCoV-19/England/ALDP-C05FA8/2020\|EPI_ISL_709304\|20 | ···················· | ···················· | ···················· | ···················· | ···················· | ···················· | |
| W | 7352 hCoV-19/England/ALDP-C06DE9/2020\|EPI_ISL_709305\|20 | ···················· | ···················· | ···················· | ···················· | ···················· | ···················· | |
| W | 7353 hCoV-19/England/ALDP-C05F4E/2020\|EPI_ISL_709308\|20 | ···················· | ···················· | ···················· | ···················· | ···················· | ···················· | |
| W | 7354 hCoV-19/England/QEUH-C146A1/2020\|EPI_ISL_709311\|20 | ···················· | ···················· | ···················· | ···················· | ···················· | ···················· | |
| W | 7355 hCoV-19/England/QEUH-C14C09/2020\|EPI_ISL_709312\|20 | ···················· | ···················· | ···················· | ···················· | ···················· | ···················· | |
| W | 7356 hCoV-19/England/ALDP-C06174/2020\|EPI_ISL_709313\|20 | ···················· | ···················· | ···················· | ···················· | ···················· | ···················· | |
| W | 7357 hCoV-19/England/ALDP-C06921/2020\|EPI_ISL_709315\|20 | ···················· | ···················· | ···················· | ···················· | ···················· | ···················· | |
| W | 7358 hCoV-19/England/ALDP-C06D43/2020\|EPI_ISL_709317\|20 | ···················· | ···················· | ···················· | ···················· | ···················· | ···················· | |
| W | 7359 hCoV-19/England/QEUH-C159F2/2020\|EPI_ISL_709320\|20 | ···················· | ···················· | ···················· | ···················· | ···················· | ···················· | |
| W | 7360 hCoV-19/England/ALDP-C06D52/2020\|EPI_ISL_709322\|20 | ···················· | ···················· | ···················· | ···················· | ···················· | ···················· | |
| W | 7361 hCoV-19/England/QEUH-C16047/2020\|EPI_ISL_709324\|20 | ···················· | ···················· | ···················· | ···················· | ···················· | ···················· | |
| W | 7362 hCoV-19/England/ALDP-C063F6/2020\|EPI_ISL_709325\|20 | ···················· | ···················· | ···················· | ···················· | ···················· | ···················· | |
| W | 7363 hCoV-19/England/ALDP-C060B3/2020\|EPI_ISL_709326\|20 | ···················· | ···················· | ···················· | ···················· | ···················· | ···················· | |
| W | 7364 hCoV-19/England/QEUH-C17F1C/2020\|EPI_ISL_709327\|20 | ···················· | ···················· | ···················· | ···················· | ···················· | ···················· | |
| W | 7365 hCoV-19/England/ALDP-C067EB/2020\|EPI_ISL_709329\|20 | ···················· | ···················· | ···················· | ···················· | ···················· | ···················· | |
| W | 7366 hCoV-19/England/QEUH-C14EE5/2020\|EPI_ISL_709336\|20 | ···················· | ···················· | ···················· | ···················· | ···················· | ···················· | |
| W | 7367 hCoV-19/England/ALDP-C05F5D/2020\|EPI_ISL_709337\|20 | ···················· | ···················· | ···················· | ···················· | ···················· | ···················· | |
| W | 7368 hCoV-19/England/ALDP-C06DF8/2020\|EPI_ISL_709340\|20 | ···················· | ···················· | ···················· | ···················· | ···················· | ···················· | |
| W | 7369 hCoV-19/England/QEUH-C159A7/2020\|EPI_ISL_709343\|20 | ···················· | ···················· | ···················· | ···················· | ···················· | ···················· | |
| W | 7370 hCoV-19/England/QEUH-C14C18/2020\|EPI_ISL_709345\|20 | ···················· | ···················· | ···················· | ···················· | ···················· | ···················· | |
| W | 7371 hCoV-19/England/ALDP-C060E0/2020\|EPI_ISL_709347\|20 | ···················· | ···················· | ···················· | ···················· | ···················· | ···················· | |
| W | 7372 hCoV-19/England/ALDP-C065F0/2020\|EPI_ISL_709348\|20 | ···················· | ·A·················· | ···················· | ···················· | ···················· | ··A················· | |
| W | 7373 hCoV-19/England/ALDP-C0646C/2020\|EPI_ISL_709349\|20 | ···················· | ···················· | ···················· | ···················· | ···················· | ···················· | |
| W | 7374 hCoV-19/England/ALDP-C064C6/2020\|EPI_ISL_709350\|20 | ···················· | ···················· | ···················· | ···················· | ···················· | ···················· | |
| W | 7375 hCoV-19/England/ALDP-C06A1F/2020\|EPI_ISL_709351\|20 | ···················· | ···················· | ···················· | ···················· | ···················· | ···················· | |
| W | 7376 hCoV-19/England/ALDP-C06693/2020\|EPI_ISL_709352\|20 | ···················· | ···················· | ···················· | ···················· | ···················· | ···················· | |
| W | 7377 hCoV-19/England/QEUH-C163B7/2020\|EPI_ISL_709353\|20 | ···················· | ···················· | ···················· | ···················· | ···················· | ···················· | |
| W | 7378 hCoV-19/England/ALDP-C06420/2020\|EPI_ISL_709356\|20 | ···················· | ···················· | ···················· | ···················· | ···················· | ···················· | |
| W | 7379 hCoV-19/England/ALDP-C06851/2020\|EPI_ISL_709357\|20 | ···················· | ···················· | ···················· | ···················· | ···················· | ···················· | |
| W | 7380 hCoV-19/England/QEUH-C1498A/2020\|EPI_ISL_709358\|20 | ···················· | ···················· | ···················· | ···················· | ···················· | ···················· | |
| W | 7381 hCoV-19/England/QEUH-C15FE1/2020\|EPI_ISL_709359\|20 | ···················· | ···················· | ···················· | ···················· | ···················· | ···················· | |
| W | 7382 hCoV-19/USA/CO-CDPHE-2004220147/2020\|EPI_ISL_71030 | ···················· | ···················· | ···················· | ···················· | ···················· | ···················· | |
| W | 7383 hCoV-19/USA/CO-CDPHE-2005220150/2020\|EPI_ISL_71030 | ···················· | ···················· | ···················· | ···················· | ···················· | ···················· | |
| W | 7384 hCoV-19/USA/CO-CDPHE-2005220166/2020\|EPI_ISL_71030 | ···················· | ···················· | ···················· | ···················· | ···················· | ···················· | |
| W | 7385 hCoV-19/USA/CO-CDPHE-2005220170/2020\|EPI_ISL_71030 | ···················· | ···················· | ···················· | ···················· | ···················· | ···················· | |
| W | 7386 hCoV-19/USA/CO-CDPHE-2005220199/2020\|EPI_ISL_71030 | ···················· | ···················· | ···················· | ···················· | ···················· | ···················· | |
| W | 7387 hCoV-19/USA/CO-CDPHE-2005220251/2020\|EPI_ISL_71030 | ···················· | ···················· | ···················· | ···················· | ···················· | ···················· | |
| W | 7388 hCoV-19/USA/CO-CDPHE-2005220253/2020\|EPI_ISL_71030 | ···················· | ···················· | ···················· | ···················· | ···················· | ···················· | |
| W | 7389 hCoV-19/USA/CO-CDPHE-2005220420/2020\|EPI_ISL_71030 | ···················· | ···················· | ···················· | ···················· | ···················· | ···················· | |
| W | 7390 hCoV-19/USA/CO-CDPHE-2005220675/2020\|EPI_ISL_71030 | ···················· | ···················· | ···················· | ···················· | ···················· | ···················· | |
| W | 7391 hCoV-19/USA/CO-CDPHE-2005220676/2020\|EPI_ISL_71031 | ···················· | ···················· | ···················· | ···················· | ···················· | ···················· | |
| W | 7392 hCoV-19/USA/CO-CDPHE-2005220677/2020\|EPI_ISL_71031 | ···················· | ···················· | ···················· | ···················· | ···················· | ···················· | |
| W | 7393 hCoV-19/USA/CO-CDPHE-2005220684/2020\|EPI_ISL_71031 | ···················· | ···················· | ···················· | ···················· | ···················· | ···················· | |
| W | 7394 hCoV-19/USA/CO-CDPHE-2005220686/2020\|EPI_ISL_71031 | ···················· | ···················· | ···················· | ···················· | ···················· | ···················· | |
| W | 7396 hCoV-19/USA/CO-CDPHE-2005270733/2020\|EPI_ISL_71031 | ···················· | ···················· | ···················· | ···················· | ···················· | ···················· | |
| W | 7397 hCoV-19/USA/CO-CDPHE-2005280131/2020\|EPI_ISL_71031 | ···················· | ···················· | ···················· | ···················· | ···················· | ···················· | |
| W | 7398 hCoV-19/USA/CO-CDPHE-2006280531/2020\|EPI_ISL_71031 | ···················· | ·A·················· | ···················· | ···················· | ···················· | ···················· | |
| W | 7399 hCoV-19/USA/CO-CDPHE-2007152317/2020\|EPI_ISL_71031 | ···················· | ···················· | ···················· | ···················· | ·NN················· | ···················· | |
| W | 7400 hCoV-19/USA/CO-CDPHE-2008021160/2020\|EPI_ISL_71031 | ···················· | ···················· | ···················· | ···················· | ···················· | ···················· | |
| W | 7401 hCoV-19/USA/CO-CDPHE-2008210131/2020\|EPI_ISL_71032 | ···················· | ···················· | ···················· | ···················· | ···················· | ···················· | |
| W | 7402 hCoV-19/USA/CO-CDPHE-2008222958/2020\|EPI_ISL_71032 | ···················· | ···················· | ···················· | ···················· | ···················· | ···················· | |
| W | 7403 hCoV-19/USA/CO-CDPHE-2008261062/2020\|EPI_ISL_71032 | ···················· | ···················· | ···················· | ···················· | ···················· | ···················· | |
| W | 7404 hCoV-19/USA/CO-CDPHE-2008281882/2020\|EPI_ISL_71032 | ···················· | ···················· | ···················· | ···················· | ···················· | ···················· | |
| W | 7405 hCoV-19/USA/CO-CDPHE-2008282003/2020\|EPI_ISL_71032 | ···················· | ···················· | ···················· | ···················· | ···················· | ···················· | |
| W | 7408 hCoV-19/USA/CO-CDPHE-2009052131/2020\|EPI_ISL_71033 | ···················· | ···················· | ···················· | ···················· | ···················· | ···················· | |
| W | 7409 hCoV-19/USA/CO-CDPHE-2009164054/2020\|EPI_ISL_71033 | ···················· | ·A·················· | ···················· | ···················· | ···················· | ···················· | |
| W | 7410 hCoV-19/USA/CO-CDPHE-2009164080/2020\|EPI_ISL_71033 | ···················· | ·A·················· | ···················· | ···················· | ···················· | ···················· | |
| W | 7411 hCoV-19/USA/CO-CDPHE-2009164105/2020\|EPI_ISL_71033 | ···················· | ·A·················· | ···················· | ···················· | ···················· | ···················· | |
| W | 7412 hCoV-19/USA/CO-CDPHE-2009164173/2020\|EPI_ISL_71033 | ···················· | ·A·················· | ···················· | ···················· | ···················· | ···················· | |
| W | 7413 hCoV-19/USA/CO-CDPHE-2009164179/2020\|EPI_ISL_71033 | ···················· | ·A·················· | ···················· | ···················· | ···················· | ···················· | |
| W | 7415 hCoV-19/USA/CO-CDPHE-2009170084/2020\|EPI_ISL_71033 | ···················· | ·A·················· | ···················· | ···················· | ···················· | ···················· | |
| W | 7416 hCoV-19/USA/CO-CDPHE-2009170087/2020\|EPI_ISL_71034 | ···················· | ·A·················· | ···················· | ···················· | ···················· | ···················· | |
| W | 7417 hCoV-19/USA/CO-CDPHE-2009170150/2020\|EPI_ISL_71034 | ···················· | ·A·················· | ···················· | ···················· | ···················· | ···················· | |
| W | 7418 hCoV-19/USA/CO-CDPHE-2009170292/2020\|EPI_ISL_71034 | ···················· | ·A·················· | ···················· | ···················· | ···················· | ···················· | |
| W | 7419 hCoV-19/USA/CO-CDPHE-2009170327/2020\|EPI_ISL_71034 | ···················· | ·A·················· | ···················· | ···················· | ···················· | ···················· | |
| W | 7420 hCoV-19/USA/CO-CDPHE-2009170329/2020\|EPI_ISL_71034 | ···················· | ·A·················· | ···················· | ···················· | ···················· | ···················· | |
| W | 7421 hCoV-19/USA/CO-CDPHE-2009170340/2020\|EPI_ISL_71034 | ···················· | ·A·················· | ···················· | ···················· | ···················· | ···················· | |
| W | 7422 hCoV-19/USA/CO-CDPHE-2009170365/2020\|EPI_ISL_71034 | ···················· | ·A·················· | ···················· | ···················· | ···················· | ···················· | |
| W | 7424 hCoV-19/USA/CO-CDPHE-2009300947/2020\|EPI_ISL_71034 | ···················· | ···················· | ···················· | ···················· | ···················· | ···················· | |
| W | 7425 hCoV-19/USA/AZ-TG389574/2020\|EPI_ISL_694700\|2020-0 | ···················· | ···················· | ···················· | ···················· | ···················· | ···················· | |
| W | 7426 hCoV-19/USA/AZ-TG392304/2020\|EPI_ISL_694701\|2020-0 | ···················· | ···················· | ···················· | ···················· | ···················· | ···················· | |
| W | 7427 hCoV-19/USA/AZ-TG392472/2020\|EPI_ISL_694702\|2020-0 | ···················· | ···················· | ···················· | ···················· | ················NN·· | ···················· | 1 |
| W | 7428 hCoV-19/USA/AZ-TG392628/2020\|EPI_ISL_694703\|2020-0 | ···················· | ···················· | ···················· | ···················· | ···················· | ···················· | |
| W | 7429 hCoV-19/USA/AZ-TG392969/2020\|EPI_ISL_694709\|2020-0 | ···················· | ···················· | ···················· | ···················· | ···················· | ···················· | |
| W | 7430 hCoV-19/USA/AZ-TG392982/2020\|EPI_ISL_694710\|2020-0 | ···················· | ···················· | ···················· | ···················· | ···················· | ···················· | |
| W | 7431 hCoV-19/USA/AZ-TG393520/2020\|EPI_ISL_694711\|2020-0 | ···················· | ·A·················· | ···················· | ···················· | ···················· | ···················· | |
| W | 7432 hCoV-19/USA/AZ-TG393553/2020\|EPI_ISL_694712\|2020-0 | ···················· | ·A·················· | ···················· | ···················· | ···················· | ···················· | |

W 7433 hCoV-19/USA/AZ-TG393888/2020|EPI_ISL_694713|2020-0 ···················· ·A·················· ···················· ···················· ···················· ····················
W 7434 hCoV-19/USA/AZ-TG393947/2020|EPI_ISL_694714|2020-0 ···················· ···················· ···················· ···················· ···················· ····················
W 7435 hCoV-19/USA/AZ-TG394115/2020|EPI_ISL_694716|2020-0 ···················· ·A·················· ···················· ···················· ···················· ····················
W 7436 hCoV-19/USA/AZ-TG395399/2020|EPI_ISL_694721|2020-0 ···················· ···················· ···················· ···················· ···················· ····················
W 7437 hCoV-19/USA/AZ-TG395813/2020|EPI_ISL_694722|2020-0 ···················· ···················· ···················· ···················· ···················· ····················
W 7438 hCoV-19/USA/AZ-TG395826/2020|EPI_ISL_694723|2020-0 ···················· ···················· ···················· ···················· ···················· ····················
W 7439 hCoV-19/USA/AZ-TG395879/2020|EPI_ISL_694724|2020-0 ···················· ···················· ···················· ···················· ···················· ····················
W 7440 hCoV-19/USA/AZ-TG395882/2020|EPI_ISL_694727|2020-0 ···················· ···················· ···················· ···················· ···················· ····················
W 7441 hCoV-19/USA/AZ-TG395887/2020|EPI_ISL_694732|2020-0 ···················· ······A············· ···················· ···················· ···················· ····················
W 7442 hCoV-19/USA/AZ-TG395894/2020|EPI_ISL_694738|2020-0 ···················· ·············A······ ···················· ···················· ···················· ····················
W 7443 hCoV-19/USA/AZ-TG395898/2020|EPI_ISL_694742|2020-0 ···················· ·············A······ ···················· ···················· ···················· ····················
W 7444 hCoV-19/USA/AZ-TG395899/2020|EPI_ISL_694743|2020-0 ···················· ·············A······ ···················· ···················· ···················· ····················
W 7445 hCoV-19/USA/AZ-TG395900/2020|EPI_ISL_694744|2020-0 ···················· ·············A······ ···················· ···················· ···················· ····················
W 7446 hCoV-19/USA/AZ-TG395902/2020|EPI_ISL_694746|2020-0 ···················· ···················· ···················· ···················· ···················· ····················
W 7447 hCoV-19/USA/AZ-TG395903/2020|EPI_ISL_694747|2020-0 ···················· ···················· ···················· ···················· ···················· ····················
W 7448 hCoV-19/USA/AZ-TG395904/2020|EPI_ISL_694748|2020-0 ···················· ···················· ···················· ···················· ···················· ····················
W 7449 hCoV-19/USA/AZ-TG395906/2020|EPI_ISL_694750|2020-0 ···················· ···················· ···················· ···················· ···················· ····················
W 7450 hCoV-19/United Arab Emirates/0658/2020|EPI_ISL_698 ···················· ···················· ···················· ···················· ···················· ····················
W 7451 hCoV-19/United Arab Emirates/0188/2020|EPI_ISL_698 ···················· ···················· ···················· ···················· ···················· ····················
W 7452 hCoV-19/United Arab Emirates/0236/2020|EPI_ISL_698 ···················· ···················· ···················· ···················· ···················· ····················
W 7453 hCoV-19/United Arab Emirates/0252/2020|EPI_ISL_698 ···················· ···················· ···················· ···················· ···················· ····················
W 7454 hCoV-19/United Arab Emirates/0072/2020|EPI_ISL_698 ···················· ···················· ···················· ···················· ···················· ····················
W 7455 hCoV-19/United Arab Emirates/0089/2020|EPI_ISL_698 ···················· ···················· ···················· ···················· ···················· ····················
W 7456 hCoV-19/United Arab Emirates/0090/2020|EPI_ISL_698 ···················· ···················· ···················· ···················· ···················· ····················
W 7457 hCoV-19/United Arab Emirates/0091/2020|EPI_ISL_698 ···················· ···················· ···················· ···················· ···················· ····················
W 7458 hCoV-19/United Arab Emirates/0092/2020|EPI_ISL_698 ···················· ···················· ···················· ···················· ···················· ····················
W 7459 hCoV-19/United Arab Emirates/0093/2020|EPI_ISL_698 ···················· ···················· ···················· ···················· ···················· ····················
W 7460 hCoV-19/United Arab Emirates/0094/2020|EPI_ISL_698 ···················· ···················· ···················· ···················· ···················· ····················
W 7461 hCoV-19/United Arab Emirates/0095/2020|EPI_ISL_698 ···················· ···················· ···················· ···················· ···················· ····················
W 7462 hCoV-19/United Arab Emirates/0097/2020|EPI_ISL_698 ···················· ···················· ···················· ···················· ···················· ····················
W 7463 hCoV-19/United Arab Emirates/0098/2020|EPI_ISL_698 ···················· ···················· ···················· ···················· ···················· ····················
W 7464 hCoV-19/United Arab Emirates/0099/2020|EPI_ISL_698 ···················· ··········T········· ···················· ···················· ···················· ····················
W 7465 hCoV-19/United Arab Emirates/0101/2020|EPI_ISL_698 ···················· ···················· ···················· ···················· ···················· ····················
W 7466 hCoV-19/United Arab Emirates/0102/2020|EPI_ISL_698 ···················· ···················· ···················· ···················· ···················· ····················
W 7467 hCoV-19/United Arab Emirates/0103/2020|EPI_ISL_698 ···················· ···················· ···················· ···················· ···················· ····················
W 7468 hCoV-19/United Arab Emirates/0104/2020|EPI_ISL_698 ···················· ···················· ···················· ···················· ···················· ····················
W 7469 hCoV-19/United Arab Emirates/0105/2020|EPI_ISL_698 ···················· ···················· ···················· ···················· ···················· ····················
W 7470 hCoV-19/United Arab Emirates/0106/2020|EPI_ISL_698 ···················· ···················· ···················· ···················· ···················· ····················
W 7471 hCoV-19/United Arab Emirates/0107/2020|EPI_ISL_698 ···················· ···················· ···················· ···················· ···················· ····················
W 7472 hCoV-19/United Arab Emirates/0108/2020|EPI_ISL_698 ···················· ···················· ···················· ···················· ···················· ····················
W 7473 hCoV-19/United Arab Emirates/0109/2020|EPI_ISL_698 ···················· ···················· ···················· ···················· ···················· ····················
W 7474 hCoV-19/England/ALDP-BBACF7/2020|EPI_ISL_703201|20 ···················· ···················· ···················· ···················· ···················· ····················
W 7475 hCoV-19/England/ALDP-BB313B/2020|EPI_ISL_703202|20 ···················· ···················· ···················· ···················· ···················· ····················
W 7476 hCoV-19/England/ALDP-BBAC9D/2020|EPI_ISL_703204|20 ···················· ···················· ···················· ···················· ···················· ····················
W 7477 hCoV-19/England/ALDP-BB3186/2020|EPI_ISL_703205|20 ···················· ···················· ···················· ···················· ···················· ····················
W 7478 hCoV-19/England/ALDP-BB5D9F/2020|EPI_ISL_703207|20 ···················· ···················· ···················· ···················· ···················· ····················
W 7479 hCoV-19/England/QEUH-BCE391/2020|EPI_ISL_704500|20 ···················· ···················· ···················· ···················· ···················· ····················
W 7480 hCoV-19/England/QEUH-BCC515/2020|EPI_ISL_704501|20 ···················· ···················· ···················· ···················· ···················· ····················
W 7481 hCoV-19/England/QEUH-BCD4F9/2020|EPI_ISL_704504|20 ···················· ···················· ···················· ···················· ···················· ····················
W 7482 hCoV-19/England/QEUH-BCCEA1/2020|EPI_ISL_704506|20 ···················· ···················· ···················· ···················· ···················· ····················
W 7483 hCoV-19/England/QEUH-BCD9FA/2020|EPI_ISL_704530|20 ···················· ···················· ···················· ···················· ···················· ····················
W 7484 hCoV-19/England/QEUH-BCBA80/2020|EPI_ISL_704531|20 ···················· ···················· ···················· ···················· ···················· ····················
W 7485 hCoV-19/England/QEUH-BCCE56/2020|EPI_ISL_704533|20 ···················· ···················· ···················· ···················· ···················· ····················
W 7486 hCoV-19/England/QEUH-BCC1A8/2020|EPI_ISL_704536|20 ···················· ···················· ···················· ···················· ···················· ····················
W 7487 hCoV-19/England/QEUH-BCE5C8/2020|EPI_ISL_704538|20 ···················· ···················· ···················· ···················· ···················· ····················
W 7489 hCoV-19/Scotland/EDB9351/2020|EPI_ISL_705801|2020- ···················· ···················· ···················· ···················· ···················· ····················
W 7490 hCoV-19/Scotland/EDB966/2020|EPI_ISL_705802|2020-0 ···················· ···················· ···················· ···················· ···················· ····················
W 7492 hCoV-19/Scotland/EDB9976/2020|EPI_ISL_705804|2020- ···················· ···················· ···················· ···················· ···················· ····················
W 7493 hCoV-19/England/LIVE-DB98E3/2020|EPI_ISL_705806|20 ···················· ···················· ···················· ···················· ···················· ····················
W 7494 hCoV-19/England/LIVE-DB9968/2020|EPI_ISL_705807|20 ···················· ···················· ···················· ···················· ···················· ····················
W 7495 hCoV-19/England/LIVE-DB9977/2020|EPI_ISL_705808|20 ···················· ···················· ···················· ···················· ···················· ····················
W 7496 hCoV-19/England/LIVE-DC18BB/2020|EPI_ISL_705809|20 ···················· ···················· ···················· ···················· ···················· ····················
W 7497 hCoV-19/England/LIVE-DC18F7/2020|EPI_ISL_705810|20 ···················· ···················· ···················· ···················· ···················· ····················
W 7498 hCoV-19/England/LIVE-DC1912/2020|EPI_ISL_705811|20 ···················· ···················· ···················· ···················· ···················· ····················
W 7499 hCoV-19/England/LIVE-DC199A/2020|EPI_ISL_705812|20 ···················· ···················· ···················· ···················· ···················· ····················
W 7500 hCoV-19/England/LIVE-DC19A9/2020|EPI_ISL_705813|20 ···················· ···················· ···················· ···················· ···················· ····················
W 7501 hCoV-19/England/LIVE-DC19E5/2020|EPI_ISL_705814|20 ···················· ···················· ···················· ···················· ···A················ ····················
W 7502 hCoV-19/England/LIVE-DC1AB5/2020|EPI_ISL_705815|20 ···················· ···················· ···················· ···················· ···················· ····················
W 7503 hCoV-19/England/LIVE-DC1BB2/2020|EPI_ISL_705816|20 ···················· ···················· ···················· ···················· ···················· ····················
W 7504 hCoV-19/England/LIVE-DC1BC1/2020|EPI_ISL_705817|20 ···················· ···················· ···················· ···················· ···················· ····················
W 7505 hCoV-19/England/LIVE-DC1BEF/2020|EPI_ISL_705818|20 ···················· ···················· ···················· ···················· ···················· ····················
W 7506 hCoV-19/England/LIVE-DC1C55/2020|EPI_ISL_705819|20 ···················· ···················· ···················· ···················· ···················· ····················
W 7507 hCoV-19/Wales/LIVE-DC1C64/2020|EPI_ISL_705820|2020 ···················· ···················· ···················· ···················· ···················· ····················
W 7508 hCoV-19/England/LIVE-DC1C73/2020|EPI_ISL_705821|20 ···················· ···················· ···················· ···················· ···················· ····················
W 7509 hCoV-19/England/LIVE-DC1D52/2020|EPI_ISL_705822|20 ···················· ···················· ···················· ···················· ···················· ····················
W 7510 hCoV-19/England/LIVE-DC1D61/2020|EPI_ISL_705823|20 ···················· ···················· ···················· ···················· ···················· ····················
W 7511 hCoV-19/England/LIVE-DC2771/2020|EPI_ISL_705824|20 ···················· ···················· ···················· ···················· ···················· ····················
W 7512 hCoV-19/England/LIVE-DC27AE/2020|EPI_ISL_705825|20 ···················· ···················· ···················· ···················· ···················· ····················
W 7513 hCoV-19/England/LIVE-DC27EA/2020|EPI_ISL_705826|20 ···················· ···················· ···················· ···················· ···A················ ····················
W 7514 hCoV-19/England/LIVE-DC286F/2020|EPI_ISL_705827|20 ···················· ···················· ···················· ···················· ···················· ····················

| | | | | | | | |
|---|---|---|---|---|---|---|---|
| W | 7515 hCoV-19/England/LIVE-DC28E7/2020\|EPI_ISL_705828\|20 | .................... | .................... | .................... | .................... | .................... | .................... |
| W | 7516 hCoV-19/England/LIVE-DC296C/2020\|EPI_ISL_705829\|20 | .................... | .................... | .................... | .................... | .................... | .................... |
| W | 7517 hCoV-19/England/LIVE-DC298A/2020\|EPI_ISL_705830\|20 | .................... | .................... | .................... | .................... | .................... | .................... |
| W | 7518 hCoV-19/England/LIVE-DC29C6/2020\|EPI_ISL_705831\|20 | .................... | .................... | .................... | .................... | .................... | .................... |
| W | 7519 hCoV-19/England/LIVE-DC2A0F/2020\|EPI_ISL_705832\|20 | .................... | .................... | .................... | .................... | .................... | .................... |
| W | 7520 hCoV-19/England/LIVE-DC2A4B/2020\|EPI_ISL_705833\|20 | .................... | .................... | .................... | .................... | .................... | .................... |
| W | 7521 hCoV-19/England/LIVE-DC2AA5/2020\|EPI_ISL_705834\|20 | .................... | .................... | .................... | .................... | .................... | .................... |
| W | 7522 hCoV-19/England/LIVE-DC2AB4/2020\|EPI_ISL_705835\|20 | .................... | .................... | .................... | .................... | .................... | .................... |
| W | 7524 hCoV-19/England/LIVE-DC2B48/2020\|EPI_ISL_705837\|20 | .................... | .................... | .................... | .................... | .................... | .................... |
| W | 7525 hCoV-19/England/LIVE-DC2B75/2020\|EPI_ISL_705838\|20 | .................... | .................... | .................... | .................... | .................... | .................... |
| W | 7526 hCoV-19/England/LIVE-DC30B1/2020\|EPI_ISL_705860\|20 | .................... | .................... | .................... | .................... | .................... | .................... |
| W | 7527 hCoV-19/England/LIVE-DC30C0/2020\|EPI_ISL_705861\|20 | .................... | .................... | .................... | .................... | .................... | .................... |
| W | 7528 hCoV-19/England/LIVE-DC30FD/2020\|EPI_ISL_705862\|20 | .................... | .................... | .................... | .................... | .................... | .................... |
| W | 7529 hCoV-19/England/LIVE-DC3127/2020\|EPI_ISL_705863\|20 | .................... | .................... | .................... | .................... | .................... | .................... |
| W | 7530 hCoV-19/England/LIVE-DC3172/2020\|EPI_ISL_705864\|20 | .................... | .................... | .................... | .................... | .................... | .................... |
| W | 7532 hCoV-19/England/LIVE-DC39F2/2020\|EPI_ISL_705866\|20 | .................... | .................... | .................... | .................... | .................... | .................... |
| W | 7535 hCoV-19/Germany/SL-SU-10452528/2020\|EPI_ISL_708001 | .................... | .................... | .................... | .................... | .................... | .................... |
| W | 7536 hCoV-19/Germany/SL-SU-10452749/2020\|EPI_ISL_708002 | .................... | .................... | .................... | .................... | .................... | .................... |
| W | 7537 hCoV-19/Norway/4602/2020\|EPI_ISL_708030\|2020-10-15 | .................... | .................... | .................... | .................... | .................... | .................... |
| W | 7538 hCoV-19/Norway/4604/2020\|EPI_ISL_708031\|2020-10-15 | .................... | .................... | .................... | .................... | .................... | .................... |
| W | 7539 hCoV-19/Norway/4629/2020\|EPI_ISL_708032\|2020-10-18 | .................... | .................... | .................... | .................... | .................... | .................... |
| W | 7540 hCoV-19/Norway/4992/2020\|EPI_ISL_708033\|2020-10-21 | .................... | .................... | .................... | .................... | .................... | .................... |
| W | 7541 hCoV-19/Norway/4995/2020\|EPI_ISL_708034\|2020-10-23 | .................... | .A.................. | .................... | .................... | .................... | ..A................. |
| W | 7542 hCoV-19/Norway/5059/2020\|EPI_ISL_708035\|2020-10-29 | .................... | .A.................. | .................... | .................... | .................... | ..A................. |
| W | 7543 hCoV-19/Norway/5091/2020\|EPI_ISL_708036\|2020-10-27 | .................... | .................... | .................... | .................... | .................... | .................... |
| W | 7544 hCoV-19/Norway/5092/2020\|EPI_ISL_708037\|2020-10-29 | .................... | .................... | .................... | .................... | .................... | .................... |
| W | 7545 hCoV-19/Norway/5094/2020\|EPI_ISL_708038\|2020-10-27 | .................... | .................... | .................... | .................... | .................... | .................... |
| W | 7546 hCoV-19/Norway/5095/2020\|EPI_ISL_708039\|2020-10-30 | .................... | .................... | .................... | .................... | .................... | .................... |
| W | 7547 hCoV-19/Norway/5121/2020\|EPI_ISL_708040\|2020-11-03 | .................... | .................... | .................... | .................... | .................... | .................... |
| W | 7548 hCoV-19/Norway/5123/2020\|EPI_ISL_708041\|2020-10-29 | .................... | .................... | .................... | .................... | .................... | .................... |
| W | 7549 hCoV-19/Norway/5165/2020\|EPI_ISL_708042\|2020-10-26 | .................... | .................... | .................... | .................... | .................... | .................... |
| W | 7550 hCoV-19/Norway/5166/2020\|EPI_ISL_708043\|2020-10-19 | .................... | .................... | .................... | .................... | .................... | .................... |
| W | 7551 hCoV-19/Norway/5171/2020\|EPI_ISL_708044\|2020-10-22 | .................... | .................... | .................... | .................... | .................... | .................... |
| W | 7552 hCoV-19/Norway/5172/2020\|EPI_ISL_708045\|2020-10-22 | .................... | .................... | .................... | .................... | .................... | .................... |
| W | 7553 hCoV-19/Norway/5182/2020\|EPI_ISL_708046\|2020-10-26 | .................... | .................... | .................... | .................... | .................... | .................... |
| W | 7554 hCoV-19/Norway/5183/2020\|EPI_ISL_708047\|2020-10-26 | .................... | .A.................. | .................... | .................... | .................... | .................... |
| W | 7555 hCoV-19/Norway/5186/2020\|EPI_ISL_708048\|2020-10-28 | .................... | .................... | .................... | .................... | .................... | .................... |
| W | 7556 hCoV-19/Norway/5209/2020\|EPI_ISL_708049\|2020-10-28 | .................... | .................... | .................... | .................... | .................... | .................... |
| W | 7557 hCoV-19/Norway/5210/2020\|EPI_ISL_708050\|2020-10-31 | .................... | .A.................. | .................... | .................... | .................... | ..A................. |
| W | 7558 hCoV-19/Norway/5211/2020\|EPI_ISL_708051\|2020-10-26 | .................... | .................... | .................... | .................... | .................... | .................... |
| W | 7559 hCoV-19/Norway/5212/2020\|EPI_ISL_708052\|2020-10-29 | .................... | .................... | .................... | .................... | .................... | .................... |
| W | 7560 hCoV-19/Norway/5217/2020\|EPI_ISL_708053\|2020-10-28 | .................... | .A.................. | .................... | .................... | .................... | ..A................. |
| W | 7561 hCoV-19/Norway/5218/2020\|EPI_ISL_708054\|2020-10-29 | .................... | .................... | .................... | .................... | .................... | .................... |
| W | 7562 hCoV-19/Norway/5239/2020\|EPI_ISL_708055\|2020-11-01 | .................... | .A.................. | .................... | .................... | .................... | ..A................. |
| W | 7563 hCoV-19/Norway/5242/2020\|EPI_ISL_708056\|2020-11-01 | .................... | .A.................. | .................... | .................... | .................... | ..A................. |
| W | 7564 hCoV-19/Norway/5323/2020\|EPI_ISL_708057\|2020-11-04 | .................... | .................... | .................... | .................... | .................... | .................... |
| W | 7565 hCoV-19/Norway/5325/2020\|EPI_ISL_708058\|2020-11-04 | .................... | .A.................. | .................... | .................... | .................... | ..A................. |
| W | 7566 hCoV-19/Norway/5327/2020\|EPI_ISL_708059\|2020-11-04 | .................... | .................... | .................... | .................... | .................... | .................... |
| W | 7567 hCoV-19/Norway/5329/2020\|EPI_ISL_708060\|2020-11-02 | .................... | .A.................. | .................... | .................... | .................... | ..A................. |
| W | 7568 hCoV-19/Norway/5330/2020\|EPI_ISL_708061\|2020-11-02 | .................... | .A.................. | .................... | .................... | .................... | ..A................. |
| W | 7569 hCoV-19/Norway/5331/2020\|EPI_ISL_708062\|2020-11-02 | .................... | .................... | .................... | .................... | .................... | .................... |
| W | 7570 hCoV-19/Norway/5333/2020\|EPI_ISL_708063\|2020-11-02 | .................... | .................... | .................... | .................... | .................... | .................... |
| W | 7571 hCoV-19/Norway/5334/2020\|EPI_ISL_708064\|2020-11-02 | .................... | .................... | .................... | .................... | .................... | .................... |
| W | 7572 hCoV-19/Norway/5366/2020\|EPI_ISL_708065\|2020-11-02 | .................... | .................... | .................... | .................... | .................... | .................... |
| W | 7573 hCoV-19/Norway/5367/2020\|EPI_ISL_708066\|2020-11-02 | .................... | .................... | .................... | .................... | .................... | .................... |
| W | 7574 hCoV-19/Norway/5369/2020\|EPI_ISL_708067\|2020-11-02 | .................... | .................... | .................... | .................... | .................... | .................... |
| W | 7575 hCoV-19/Norway/5371/2020\|EPI_ISL_708068\|2020-11-02 | .................... | .................... | .................... | .................... | .................... | .................... |
| W | 7576 hCoV-19/Norway/5373/2020\|EPI_ISL_708069\|2020-10-28 | ........T........... | .A.................. | .................... | .................... | .................... | .................... |
| W | 7577 hCoV-19/Norway/5377/2020\|EPI_ISL_708070\|2020-10-26 | .................... | .................... | .................... | .................... | .................... | .................... |
| W | 7578 hCoV-19/Norway/5380/2020\|EPI_ISL_708071\|2020-10-28 | .................... | .A.................. | .................... | .................... | .................... | ..A................. |
| W | 7579 hCoV-19/Norway/5386/2020\|EPI_ISL_708072\|2020-10-28 | .................... | .................... | .................... | .................... | .................... | .................... |
| W | 7580 hCoV-19/Norway/5387/2020\|EPI_ISL_708073\|2020-10-28 | .................... | .................... | .................... | .................... | .................... | .................... |
| W | 7581 hCoV-19/Norway/5393/2020\|EPI_ISL_708074\|2020-10-27 | .................... | .................... | .................... | .................... | .................... | .................... |
| W | 7582 hCoV-19/Norway/5394/2020\|EPI_ISL_708075\|2020-11-04 | .................... | .................... | .................... | .................... | .................... | .................... |
| W | 7583 hCoV-19/Norway/5396/2020\|EPI_ISL_708076\|2020-11-05 | .................... | .................... | .................... | .................... | .................... | .................... |
| W | 7584 hCoV-19/Norway/5397/2020\|EPI_ISL_708077\|2020-11-04 | .................... | .A.................. | .................... | .................... | .................... | ..A................. |
| W | 7585 hCoV-19/Norway/5399/2020\|EPI_ISL_708078\|2020-11-02 | .................... | .................... | .................... | .................... | .................... | .................... |
| W | 7586 hCoV-19/Norway/5401/2020\|EPI_ISL_708079\|2020-10-26 | .................... | .A.................. | .................... | .................... | .................... | .................... |
| W | 7587 hCoV-19/Norway/5404/2020\|EPI_ISL_708080\|2020-11-03 | .................... | .................... | .................... | .................... | .................... | .................... |
| W | 7588 hCoV-19/Norway/5410/2020\|EPI_ISL_708081\|2020-11-02 | .................... | .................... | .................... | .................... | .................... | .................... |
| W | 7589 hCoV-19/Norway/5415/2020\|EPI_ISL_708082\|2020-10-28 | .................... | .A.................. | .................... | .................... | .................... | .................... |
| W | 7590 hCoV-19/Norway/5416/2020\|EPI_ISL_708083\|2020-10-28 | .................... | .A.................. | .................... | .................... | .................... | .................... |
| W | 7591 hCoV-19/Norway/5417/2020\|EPI_ISL_708084\|2020-10-29 | .................... | .A.................. | .................... | .................... | .................... | .................... |
| W | 7592 hCoV-19/Norway/5418/2020\|EPI_ISL_708085\|2020-10-28 | .................... | .A.................. | .................... | .................... | .................... | .................... |
| W | 7593 hCoV-19/Norway/5419/2020\|EPI_ISL_708086\|2020-11-03 | .................... | .................... | .................... | .................... | .................... | .................... |
| W | 7594 hCoV-19/Norway/5421/2020\|EPI_ISL_708087\|2020-11-03 | .................... | .................... | .................... | .................... | .................... | .................... |
| W | 7595 hCoV-19/Norway/5422/2020\|EPI_ISL_708088\|2020-11-03 | .................... | .................... | .................... | .................... | .................... | .................... |
| W | 7596 hCoV-19/Norway/5423/2020\|EPI_ISL_708089\|2020-11-03 | .................... | .................... | .................... | .................... | .................... | .................... |
| W | 7597 hCoV-19/Norway/5425/2020\|EPI_ISL_708090\|2020-11-03 | .................... | .................... | .................... | .................... | .................... | .................... |
| W | 7598 hCoV-19/Norway/5429/2020\|EPI_ISL_708091\|2020-10-26 | .................... | .................... | .................... | .................... | .................... | .................... |

W 7599 hCoV-19/Norway/5432/2020|EPI_ISL_708092|2020-11-03 ···················· ···················· ····················· ···················· ···················· ····················
W 7600 hCoV-19/Norway/5433/2020|EPI_ISL_708093|2020-10-30 ···················· ···················· ····················· ···················· ···················· ····················
W 7601 hCoV-19/Norway/5434/2020|EPI_ISL_708094|2020-11-02 ···················· ···················· ····················· ···················· ···················· ····················
W 7602 hCoV-19/Norway/5436/2020|EPI_ISL_708095|2020-10-27 ···················· ·A·················· ····················· ···················· ···················· ··A·················
W 7603 hCoV-19/Norway/5452/2020|EPI_ISL_708096|2020-11-03 ···················· ·A·················· ····················· ···················· ···················· ··A·················
W 7604 hCoV-19/Norway/5455/2020|EPI_ISL_708097|2020-11-03 ···················· ···················· ····················· ···················· ···················· ····················
W 7605 hCoV-19/Norway/5459/2020|EPI_ISL_708098|2020-10-28 ···················· ···················· ····················· ···················· ···················· ····················
W 7606 hCoV-19/Norway/5460/2020|EPI_ISL_708099|2020-10-28 ···················· ···················· ····················· ···················· ···················· ····················
W 7607 hCoV-19/England/QEUH-C14577/2020|EPI_ISL_709332|20 ···················· ···················· ····················· ···················· ···················· ····················
W 7608 hCoV-19/England/QEUH-C145C2/2020|EPI_ISL_709334|20 ···················· ···················· ····················· ···················· ···················· ····················
W 7609 hCoV-19/England/ALDP-C062AE/2020|EPI_ISL_709339|20 ···················· ···················· ····················· ···················· ···················· ····················
W 7610 hCoV-19/England/QEUH-C14B48/2020|EPI_ISL_709360|20 ···················· ·A·················· ····················· ···················· ···················· ··A·················
W 7611 hCoV-19/England/ALDP-C06183/2020|EPI_ISL_709361|20 ···················· ···················· ····················· ···················· ···················· ····················
W 7612 hCoV-19/England/ALDP-C0647B/2020|EPI_ISL_709362|20 ···················· ···················· ····················· ···················· ···················· ····················
W 7613 hCoV-19/England/ALDP-C0643F/2020|EPI_ISL_709363|20 ···················· ···················· ····················· ···················· ···················· ····················
W 7614 hCoV-19/England/QEUH-C149F3/2020|EPI_ISL_709364|20 ···················· ···················· ····················· ···················· ···················· ····················
W 7615 hCoV-19/England/QEUH-C14999/2020|EPI_ISL_709365|20 ···················· ·A·················· ····················· ···················· ···················· ····················
W 7616 hCoV-19/England/QEUH-C145B3/2020|EPI_ISL_709367|20 ···················· ···················· ····················· ···················· ···················· ····················
W 7617 hCoV-19/England/ALDP-C0600E/2020|EPI_ISL_709369|20 ···················· ···················· ····················· ···················· ···················· ····················
W 7618 hCoV-19/England/ALDP-C069F4/2020|EPI_ISL_709371|20 ···················· ···················· ····················· ···················· ···················· ····················
W 7619 hCoV-19/England/ALDP-C06077/2020|EPI_ISL_709372|20 ···················· ···················· ····················· ···················· ···················· ····················
W 7620 hCoV-19/England/QEUH-C17EE2/2020|EPI_ISL_709373|20 ···················· ···················· ····················· ···················· ···················· ····················
W 7621 hCoV-19/England/ALDP-C05FE4/2020|EPI_ISL_709374|20 ···················· ···················· ····················· ···················· ···················· ····················
W 7622 hCoV-19/England/ALDP-C05FB7/2020|EPI_ISL_709375|20 ···················· ·A·················· ····················· ···················· ···················· ··A·················
W 7623 hCoV-19/England/QEUH-C15F87/2020|EPI_ISL_709376|20 ···················· ···················· ····················· ···················· ···················· ····················
W 7624 hCoV-19/England/ALDP-C06D8F/2020|EPI_ISL_709379|20 ···················· ···················· ····················· ···················· ···················· ····················
W 7625 hCoV-19/England/QEUH-C15F96/2020|EPI_ISL_709380|20 ···················· ···················· ····················· ···················· ···················· ····················
W 7626 hCoV-19/England/ALDP-C06763/2020|EPI_ISL_709382|20 ···················· ···················· ····················· ···················· ···················· ····················
W 7627 hCoV-19/England/QEUH-C14ED6/2020|EPI_ISL_709383|20 ···················· ···················· ·········T··········· ···················· ···················· ····················
W 7628 hCoV-19/England/QEUH-C16029/2020|EPI_ISL_709384|20 ···················· ···················· ····················· ···················· ···················· ····················
W 7629 hCoV-19/England/ALDP-C0603B/2020|EPI_ISL_709386|20 ···················· ···················· ····················· ···················· ···················· ····················
W 7630 hCoV-19/England/ALDP-C06D9E/2020|EPI_ISL_709390|20 ···················· ···················· ····················· ···················· ···················· ····················
W 7631 hCoV-19/England/QEUH-C1601A/2020|EPI_ISL_709392|20 ···················· ···················· ····················· ···················· ···················· ····················
W 7632 hCoV-19/England/ALDP-C0644E/2020|EPI_ISL_709393|20 ···················· ···················· ····················· ···················· ···················· ····················
W 7633 hCoV-19/England/QEUH-C146B0/2020|EPI_ISL_709395|20 ···················· ···················· ····················· ···················· ···················· ····················
W 7634 hCoV-19/England/QEUH-C14B66/2020|EPI_ISL_709396|20 ···················· ···················· ····················· ···················· ···················· ····················
W 7635 hCoV-19/England/ALDP-C0650F/2020|EPI_ISL_709397|20 ···················· ···················· ····················· ···················· ···················· ····················
W 7636 hCoV-19/England/QEUH-C14E4F/2020|EPI_ISL_709399|20 ···················· ···················· ····················· ···················· ···················· ····················
W 7638 hCoV-19/USA/CO-CDPHE-2008063525/2020|EPI_ISL_71032 ···················· ···················· ····················· ···················· ···················· ····················
W 7639 hCoV-19/USA/CO-CDPHE-2008081584/2020|EPI_ISL_71032 ···················· ·A·················· ····················· ···················· ···················· ····················
W 7640 hCoV-19/USA/CO-CDPHE-2008170978/2020|EPI_ISL_71032 ···················· ·A·················· ····················· ···················· ···················· ····················
W 7642 hCoV-19/USA/CO-CDPHE-2010112562/2020|EPI_ISL_71034 ···················· ···················· ····················· ···················· ···················· ····················
W 7643 hCoV-19/USA/CO-CDPHE-2010112598/2020|EPI_ISL_71035 ···················· ···················· ····················· ···················· ···················· ····················
W 7644 hCoV-19/USA/CO-CDPHE-2010120393/2020|EPI_ISL_71035 ···················· ···················· ····················· ···················· ···················· ····················
W 7645 hCoV-19/USA/CO-CDPHE-2010130202/2020|EPI_ISL_71035 ···················· ···················· ····················· ···················· ···················· ····················
W 7647 hCoV-19/USA/CO-CDPHE-2010191275/2020|EPI_ISL_71035 ···················· ·A·················· ····················· ···················· ···················· ····················
W 7648 hCoV-19/USA/CO-CDPHE-2010200648/2020|EPI_ISL_71035 ···················· ···················· ····················· ···················· ···················· ····················
W 7649 hCoV-19/USA/CO-CDPHE-2010200764/2020|EPI_ISL_71035 ···················· ···················· ····················· ···················· ···················· ····················
W 7650 hCoV-19/USA/CO-CDPHE-2010200815/2020|EPI_ISL_71035 ···················· ···················· ····················· ···················· ···················· ····················
W 7651 hCoV-19/USA/CO-CDPHE-2010201090/2020|EPI_ISL_71035 ···················· ···················· ····················· ···················· ···················· ····················
W 7652 hCoV-19/USA/CO-CDPHE-2010230051/2020|EPI_ISL_71035 ···················· ···················· ····················· ···················· ···················· ····················
W 7653 hCoV-19/USA/CO-CDPHE-2010280186/2020|EPI_ISL_71036 ···················· ···················· ····················· ···················· ···················· ····················
W 7654 hCoV-19/USA/CO-CDPHE-2010280189/2020|EPI_ISL_71036 ···················· ···················· ····················· ···················· ···················· ····················
W 7656 hCoV-19/USA/CO-CDPHE-2010280209/2020|EPI_ISL_71036 ···················· ···················· ····················· ···················· ···················· ····················
W 7657 hCoV-19/USA/CO-CDPHE-2010280211/2020|EPI_ISL_71036 ···················· ···················· ····················· ···················· ···················· ····················
W 7658 hCoV-19/USA/CO-CDPHE-2010280212/2020|EPI_ISL_71036 ···················· ···················· ····················· ···················· ···················· ····················
W 7659 hCoV-19/USA/CO-CDPHE-2010280214/2020|EPI_ISL_71036 ···················· ···················· ····················· ···················· ···················· ····················
W 7660 hCoV-19/USA/CO-CDPHE-2010280216/2020|EPI_ISL_71036 ···················· ···················· ····················· ···················· ···················· ····················
W 7662 hCoV-19/USA/CO-CDPHE-2011020252/2020|EPI_ISL_71036 ···················· ···················· ····················· ···················· ···················· ····················
W 7663 hCoV-19/USA/CO-CDPHE-2011020263/2020|EPI_ISL_71037 ···················· ···················· ····················· ···················· ···················· ····················
W 7664 hCoV-19/USA/CO-CDPHE-2011020274/2020|EPI_ISL_71037 ···················· ···················· ····················· ···················· ···················· ····················
W 7665 hCoV-19/USA/CO-CDPHE-2011020311/2020|EPI_ISL_71037 ···················· ···················· ····················· ···················· ···················· ····················
W 7666 hCoV-19/USA/CO-CDPHE-2011020331/2020|EPI_ISL_71037 ···················· ···················· ····················· ···················· ···················· ····················
W 7667 hCoV-19/USA/TX-DSHS-1760/2020|EPI_ISL_710385|2020- ···················· ···················· ····················· ···················· ···················· ····················
W 7668 hCoV-19/USA/TX-DSHS-1735/2020|EPI_ISL_710387|2020- ···················· ···················· ····················· ···················· ···················· ····················
W 7669 hCoV-19/USA/TX-DSHS-1729/2020|EPI_ISL_710388|2020- ···················· ···················· ····················· ···················· ···················· ····················
W 7670 hCoV-19/USA/TX-DSHS-1730/2020|EPI_ISL_710389|2020- ···················· ···················· ····················· ···················· ···················· ····················
W 7671 hCoV-19/USA/TX-DSHS-1732/2020|EPI_ISL_710390|2020- ···················· ···················· ····················· ···················· ···················· ····················
W 7672 hCoV-19/USA/TX-DSHS-1734/2020|EPI_ISL_710391|2020- ···················· ···················· ····················· ···················· ···················· ····················
W 7673 hCoV-19/USA/TX-DSHS-1736/2020|EPI_ISL_710392|2020- ···················· ·············A······ ····················· ···················· ···················· ····················
W 7674 hCoV-19/USA/TX-DSHS-1737/2020|EPI_ISL_710393|2020- ···················· ···················· ····················· ···················· ···················· ····················
W 7675 hCoV-19/USA/TX-DSHS-1738/2020|EPI_ISL_710394|2020- ···················· ···················· ····················· ···················· ···················· ····················
W 7676 hCoV-19/USA/TX-DSHS-1739/2020|EPI_ISL_710395|2020- ···················· ···················· ····················· ···················· ···················· ····················
W 7677 hCoV-19/USA/TX-DSHS-1742/2020|EPI_ISL_710396|2020- ···················· ···················· ····················· ···················· ···················· ····················
W 7678 hCoV-19/USA/TX-DSHS-1744/2020|EPI_ISL_710397|2020- ···················· ···················· ····················· ···················· ···················· ····················
W 7679 hCoV-19/USA/TX-DSHS-1746/2020|EPI_ISL_710398|2020- ···················· ···················· ····················· ···················· ···················· ····················
W 7680 hCoV-19/USA/TX-DSHS-1747/2020|EPI_ISL_710399|2020- ···················· ···················· ····················· ···················· ···················· ····················
W 7681 hCoV-19/United Arab Emirates/0290/2020|EPI_ISL_698 ···················· ···················· ····················· ···················· ···················· ····················
W 7682 hCoV-19/United Arab Emirates/0536/2020|EPI_ISL_698 ···················· ···················· ····················· ···················· ···················· ····················
W 7683 hCoV-19/United Arab Emirates/0877/2020|EPI_ISL_698 ···················· ···················· ····················· ···················· ···················· ····················

| | | | | | | | | |
|---|---|---|---|---|---|---|---|---|
| W | 7684 | hCoV-19/United Arab Emirates/0305/2020\|EPI_ISL_698 | .................... | .................... | .................... | .................... | .................... | .................... |
| W | 7685 | hCoV-19/United Arab Emirates/0001/2020\|EPI_ISL_698 | .................... | .................... | .................... | .................... | .................... | .................... |
| W | 7686 | hCoV-19/United Arab Emirates/0002/2020\|EPI_ISL_698 | .................... | .................... | .................... | .................... | .................... | .................... |
| W | 7687 | hCoV-19/United Arab Emirates/0003/2020\|EPI_ISL_698 | .................... | .................... | .................... | .................... | .................... | .................... |
| W | 7688 | hCoV-19/United Arab Emirates/0004/2020\|EPI_ISL_698 | .................... | .................... | .................... | .................... | .................... | .................... |
| W | 7689 | hCoV-19/United Arab Emirates/0005/2020\|EPI_ISL_698 | .................... | .................... | .................... | .................... | .................... | .................... |
| W | 7690 | hCoV-19/United Arab Emirates/0006/2020\|EPI_ISL_698 | .................... | .................... | .................... | .................... | .................... | .................... |
| W | 7691 | hCoV-19/United Arab Emirates/0009/2020\|EPI_ISL_698 | .................... | .................... | .................... | .................... | .................... | .................... |
| W | 7692 | hCoV-19/United Arab Emirates/0010/2020\|EPI_ISL_698 | .................... | .................... | .................... | .................... | .................... | .................... |
| W | 7693 | hCoV-19/United Arab Emirates/0011/2020\|EPI_ISL_698 | .................... | .................... | .................... | .................... | .................... | .................... |
| W | 7694 | hCoV-19/United Arab Emirates/0012/2020\|EPI_ISL_698 | .................... | .................... | .................... | .................... | .................... | .................... |
| W | 7695 | hCoV-19/United Arab Emirates/0013/2020\|EPI_ISL_698 | .................... | .................... | .................... | .................... | .................... | .................... |
| W | 7696 | hCoV-19/United Arab Emirates/0014/2020\|EPI_ISL_698 | .................... | .................... | .................... | .................... | .................... | .................... |
| W | 7697 | hCoV-19/United Arab Emirates/0015/2020\|EPI_ISL_698 | .................... | .................... | .................... | .................... | .................... | .................... |
| W | 7698 | hCoV-19/United Arab Emirates/0016/2020\|EPI_ISL_698 | .................... | .................... | .................... | .................... | .................... | .................... |
| W | 7699 | hCoV-19/United Arab Emirates/0017/2020\|EPI_ISL_698 | .................... | .................... | .................... | .................... | .................... | .................... |
| W | 7700 | hCoV-19/United Arab Emirates/0018/2020\|EPI_ISL_698 | .................... | .................... | .................... | .................... | .................... | .................... |
| W | 7701 | hCoV-19/United Arab Emirates/0019/2020\|EPI_ISL_698 | .................... | .................... | .................... | .................... | .................... | .................... |
| W | 7702 | hCoV-19/United Arab Emirates/0020/2020\|EPI_ISL_698 | .................... | .................... | .................... | .................... | .................... | .................... |
| W | 7703 | hCoV-19/United Arab Emirates/0021/2020\|EPI_ISL_698 | .................... | .................... | .................... | .................... | .................... | .................... |
| W | 7704 | hCoV-19/United Arab Emirates/0022/2020\|EPI_ISL_698 | .................... | .................... | .................... | .................... | .................... | .................... |
| W | 7705 | hCoV-19/United Arab Emirates/0023/2020\|EPI_ISL_698 | .................... | .................... | .................... | .................... | .................... | .................... |
| W | 7706 | hCoV-19/United Arab Emirates/0025/2020\|EPI_ISL_698 | .................... | .................... | .................... | .................... | .................... | .................... |
| W | 7707 | hCoV-19/United Arab Emirates/0026/2020\|EPI_ISL_698 | .................... | .................... | .................... | .................... | .................... | .................... |
| W | 7708 | hCoV-19/United Arab Emirates/0027/2020\|EPI_ISL_698 | .................... | .................... | .................... | .................... | .................... | .................... |
| W | 7709 | hCoV-19/United Arab Emirates/0028/2020\|EPI_ISL_698 | .................... | .................... | .................... | .................... | .................... | .................... |
| W | 7710 | hCoV-19/United Arab Emirates/0029/2020\|EPI_ISL_698 | .................... | .................... | .................... | .................... | .................... | .................... |
| W | 7711 | hCoV-19/United Arab Emirates/0030/2020\|EPI_ISL_698 | .................... | .................... | .................... | .................... | .................... | .................... |
| W | 7712 | hCoV-19/United Arab Emirates/0031/2020\|EPI_ISL_698 | .................... | .................... | .................... | .................... | .................... | .................... |
| W | 7713 | hCoV-19/United Arab Emirates/0032/2020\|EPI_ISL_698 | .................... | .................... | .................... | .................... | .................... | .................... |
| W | 7714 | hCoV-19/United Arab Emirates/0033/2020\|EPI_ISL_698 | .................... | .................... | .................... | .................... | .................... | .................... |
| W | 7715 | hCoV-19/United Arab Emirates/0034/2020\|EPI_ISL_698 | .................... | .................... | .................... | .................... | .................... | .................... |
| W | 7716 | hCoV-19/United Arab Emirates/0035/2020\|EPI_ISL_698 | .................... | .................... | .................... | .................... | .................... | .................... |
| W | 7717 | hCoV-19/United Arab Emirates/0036/2020\|EPI_ISL_698 | .................... | .................... | .................... | .................... | .................... | .................... |
| W | 7718 | hCoV-19/United Arab Emirates/0037/2020\|EPI_ISL_698 | .................... | .................... | .................... | .................... | .................... | .................... |
| W | 7719 | hCoV-19/United Arab Emirates/0038/2020\|EPI_ISL_698 | .................... | .................... | .................... | .................... | .................... | .................... |
| W | 7720 | hCoV-19/United Arab Emirates/0039/2020\|EPI_ISL_698 | .................... | .................... | .................... | .................... | .................... | .................... |
| W | 7721 | hCoV-19/United Arab Emirates/0040/2020\|EPI_ISL_698 | .................... | .................... | .................... | .................... | .................... | .................... |
| W | 7722 | hCoV-19/United Arab Emirates/0041/2020\|EPI_ISL_698 | .................... | .................... | .................... | .................... | .................... | .................... |
| W | 7723 | hCoV-19/United Arab Emirates/0042/2020\|EPI_ISL_698 | .................... | .................... | .................... | .................... | .................... | .................... |
| W | 7724 | hCoV-19/United Arab Emirates/0043/2020\|EPI_ISL_698 | .................... | .................... | .................... | .................... | .................... | .................... |
| W | 7725 | hCoV-19/United Arab Emirates/0044/2020\|EPI_ISL_698 | .................... | .................... | .................... | .................... | .................... | .................... |
| W | 7726 | hCoV-19/United Arab Emirates/0045/2020\|EPI_ISL_698 | .................... | .................... | .................... | .................... | .................... | .................... |
| W | 7727 | hCoV-19/United Arab Emirates/0047/2020\|EPI_ISL_698 | .................... | .................... | .................... | .................... | .................... | .................... |
| W | 7728 | hCoV-19/United Arab Emirates/0048/2020\|EPI_ISL_698 | .................... | .................... | .................... | .................... | .................... | .................... |
| W | 7729 | hCoV-19/United Arab Emirates/0049/2020\|EPI_ISL_698 | .................... | .................... | .................... | .................... | .................... | .................... |
| W | 7730 | hCoV-19/United Arab Emirates/0050/2020\|EPI_ISL_698 | .................... | .................... | .................... | .................... | .................... | .................... |
| W | 7731 | hCoV-19/United Arab Emirates/0051/2020\|EPI_ISL_698 | .................... | .................... | .................... | .................... | .................... | .................... |
| W | 7732 | hCoV-19/United Arab Emirates/0052/2020\|EPI_ISL_698 | .................... | .................... | .................... | .................... | .................... | .................... |
| W | 7733 | hCoV-19/United Arab Emirates/0053/2020\|EPI_ISL_698 | .................... | .................... | .................... | .................... | .................... | .................... |
| W | 7734 | hCoV-19/United Arab Emirates/0054/2020\|EPI_ISL_698 | .................... | .................... | .................... | .................... | .................... | .................... |
| W | 7735 | hCoV-19/United Arab Emirates/0055/2020\|EPI_ISL_698 | .................... | .................... | .................... | .................... | .................... | .................... |
| W | 7736 | hCoV-19/United Arab Emirates/0056/2020\|EPI_ISL_698 | .................... | .................... | .................... | .................... | .................... | .................... |
| W | 7737 | hCoV-19/United Arab Emirates/0057/2020\|EPI_ISL_698 | .................... | .................... | .................... | .................... | .................... | .................... |
| W | 7738 | hCoV-19/United Arab Emirates/0058/2020\|EPI_ISL_698 | .................... | .................... | .................... | .................... | .................... | .................... |
| W | 7739 | hCoV-19/United Arab Emirates/0059/2020\|EPI_ISL_698 | .................... | .................... | .................... | .................... | .................... | .................... |
| W | 7740 | hCoV-19/United Arab Emirates/0060/2020\|EPI_ISL_698 | .................... | .................... | .................... | .................... | .................... | .................... |
| W | 7741 | hCoV-19/United Arab Emirates/0061/2020\|EPI_ISL_698 | .................... | .................... | .................... | .................... | .................... | .................... |
| W | 7742 | hCoV-19/United Arab Emirates/0062/2020\|EPI_ISL_698 | .................... | .................... | .................... | .................... | .................... | .................... |
| W | 7743 | hCoV-19/United Arab Emirates/0063/2020\|EPI_ISL_698 | .................... | .................... | .................... | .................... | .................... | .................... |
| W | 7744 | hCoV-19/United Arab Emirates/0064/2020\|EPI_ISL_698 | .................... | .................... | .................... | .................... | .................... | .................... |
| W | 7745 | hCoV-19/United Arab Emirates/0065/2020\|EPI_ISL_698 | .................... | .................... | .................... | .................... | .................... | .................... |
| W | 7746 | hCoV-19/United Arab Emirates/0066/2020\|EPI_ISL_698 | .................... | .................... | .................... | .................... | .................... | .................... |
| W | 7747 | hCoV-19/United Arab Emirates/0067/2020\|EPI_ISL_698 | .................... | .................... | .................... | .................... | .................... | .................... |
| W | 7748 | hCoV-19/United Arab Emirates/0068/2020\|EPI_ISL_698 | .................... | .................... | .................... | .................... | .................... | .................... |
| W | 7749 | hCoV-19/United Arab Emirates/0069/2020\|EPI_ISL_698 | .................... | .................... | .................... | .................... | .................... | .................... |
| W | 7750 | hCoV-19/United Arab Emirates/0070/2020\|EPI_ISL_698 | .................... | .................... | .................... | .................... | .................... | .................... |
| W | 7751 | hCoV-19/United Arab Emirates/0071/2020\|EPI_ISL_698 | .................... | .................... | .................... | .................... | .................... | .................... |
| W | 7752 | hCoV-19/England/ALDP-BB8F69/2020\|EPI_ISL_703210\|20 | .................... | .................... | .................... | .................... | .................... | .................... |
| W | 7753 | hCoV-19/England/ALDP-BB5791/2020\|EPI_ISL_703212\|20 | .................... | .................... | .................... | .................... | .................... | .................... |
| W | 7754 | hCoV-19/England/ALDP-BB98F4/2020\|EPI_ISL_703215\|20 | .................... | .................... | .................... | .................... | .................... | .................... |
| W | 7755 | hCoV-19/England/ALDP-BB2B5C/2020\|EPI_ISL_703217\|20 | .................... | .................... | .................... | .................... | .................... | .................... |
| W | 7756 | hCoV-19/England/ALDP-BB9900/2020\|EPI_ISL_703220\|20 | .................... | .................... | .................... | .................... | .................... | .................... |
| W | 7757 | hCoV-19/England/ALDP-BB280A/2020\|EPI_ISL_703221\|20 | .................... | .................... | .................... | .................... | .................... | .................... |
| W | 7758 | hCoV-19/England/ALDP-BB8C26/2020\|EPI_ISL_703224\|20 | .................... | .................... | .................... | .................... | .................... | .................... |
| W | 7759 | hCoV-19/England/ALDP-BB8E99/2020\|EPI_ISL_703226\|20 | .................... | .................... | .................... | .................... | .................... | .................... |
| W | 7760 | hCoV-19/England/ALDP-BB8EA8/2020\|EPI_ISL_703231\|20 | .................... | .................... | .................... | .................... | .................... | .................... |
| W | 7761 | hCoV-19/England/ALDP-BB9CAD/2020\|EPI_ISL_703232\|20 | .................... | .................... | .................... | .................... | .................... | .................... |
| W | 7762 | hCoV-19/England/ALDP-BB31B3/2020\|EPI_ISL_703236\|20 | .................... | .................... | .................... | .................... | .................... | .................... |
| W | 7763 | hCoV-19/England/ALDP-BB56A3/2020\|EPI_ISL_703237\|20 | .................... | .................... | .................... | .................... | .................... | .................... |

| | | | | | | | | |
|---|---|---|---|---|---|---|---|---|
| W | 7764 | hCoV-19/England/ALDP-BB5306/2020\|EPI_ISL_703239\|20 | ···················· | ···················· | ···················· | ···················· | ···················· | ···················· |
| W | 7765 | hCoV-19/England/ALDP-BB55B5/2020\|EPI_ISL_703242\|20 | ···················· | ···················· | ···················· | ···················· | ···················· | ···················· |
| W | 7766 | hCoV-19/England/ALDP-BCA896/2020\|EPI_ISL_703247\|20 | ···················· | ···················· | ···················· | ···················· | ···················· | ···················· |
| W | 7767 | hCoV-19/England/ALDP-BCA1C8/2020\|EPI_ISL_703248\|20 | ···················· | ···················· | ···················· | ···················· | ·····T·············· | ···················· |
| W | 7768 | hCoV-19/England/ALDP-BCA702/2020\|EPI_ISL_703250\|20 | ···················· | ···················· | ···················· | ···················· | ···················· | ···················· |
| W | 7769 | hCoV-19/England/ALDP-BCAA63/2020\|EPI_ISL_703252\|20 | ···················· | ···················· | ···················· | ···················· | ···················· | ···················· |
| W | 7770 | hCoV-19/England/ALDP-BCA456/2020\|EPI_ISL_703253\|20 | ···················· | ···················· | ···················· | ···················· | ···················· | ···················· |
| W | 7771 | hCoV-19/England/ALDP-BCA1E6/2020\|EPI_ISL_703255\|20 | ···················· | ···················· | ···················· | ···················· | ···················· | ···················· |
| W | 7772 | hCoV-19/England/ALDP-BCA4A1/2020\|EPI_ISL_703257\|20 | ···················· | ···················· | ···················· | ···················· | ···················· | ···················· |
| W | 7773 | hCoV-19/England/QEUH-BCBBBA/2020\|EPI_ISL_704508\|20 | ···················· | ···················· | ···················· | ···················· | ···················· | ···················· |
| W | 7774 | hCoV-19/England/QEUH-BCBD3C/2020\|EPI_ISL_704511\|20 | ···················· | ···················· | ···················· | ···················· | ···················· | ···················· |
| W | 7775 | hCoV-19/England/QEUH-BCD1A7/2020\|EPI_ISL_704512\|20 | ···················· | ···················· | ···················· | ···················· | ···················· | ···················· |
| W | 7776 | hCoV-19/England/QEUH-BCB516/2020\|EPI_ISL_704514\|20 | ···················· | ···················· | ···················· | ···················· | ···················· | ···················· |
| W | 7777 | hCoV-19/England/QEUH-BCBADB/2020\|EPI_ISL_704523\|20 | ···················· | ···················· | ···················· | ···················· | ···················· | ···················· |
| W | 7778 | hCoV-19/England/QEUH-BCCF53/2020\|EPI_ISL_704525\|20 | ···················· | ···················· | ···················· | ···················· | ···················· | ···················· |
| W | 7779 | hCoV-19/England/QEUH-BCD963/2020\|EPI_ISL_704527\|20 | ···················· | ···················· | ···················· | ···················· | ···················· | ···················· |
| W | 7780 | hCoV-19/England/QEUH-BCE8FC/2020\|EPI_ISL_704539\|20 | ···················· | ···················· | ···················· | ···················· | ···················· | ···················· |
| W | 7781 | hCoV-19/England/QEUH-BCE8A1/2020\|EPI_ISL_704541\|20 | ···················· | ···················· | ···················· | ···················· | ···················· | ···················· |
| W | 7782 | hCoV-19/England/QEUH-BCC4EB/2020\|EPI_ISL_704542\|20 | ···················· | ·A·················· | ···················· | ···················· | ···················· | ··A················· |
| W | 7783 | hCoV-19/England/QEUH-BCCADA/2020\|EPI_ISL_704544\|20 | ···················· | ···················· | ···················· | ···················· | ···················· | ···················· |
| W | 7784 | hCoV-19/England/QEUH-BCE971/2020\|EPI_ISL_704547\|20 | ···················· | ···················· | ···················· | ···················· | ···················· | ···················· |
| W | 7785 | hCoV-19/England/QEUH-BCE56E/2020\|EPI_ISL_704549\|20 | ···················· | ···················· | ···················· | ···················· | ·····T·············· | ···················· |
| W | 7786 | hCoV-19/England/QEUH-BCDA15/2020\|EPI_ISL_704550\|20 | ···················· | ···················· | ···················· | ···················· | ···················· | ···················· |
| W | 7787 | hCoV-19/England/QEUH-BCC20F/2020\|EPI_ISL_704552\|20 | ···················· | ···················· | ···················· | ···················· | ···················· | ···················· |
| W | 7788 | hCoV-19/England/QEUH-BCE2C1/2020\|EPI_ISL_704556\|20 | ···················· | ···················· | ···················· | ···················· | ···················· | ···················· |
| W | 7789 | hCoV-19/England/QEUH-BCBDC3/2020\|EPI_ISL_704558\|20 | ···················· | ···················· | ···················· | ···················· | ·····T·············· | ···················· |
| W | 7790 | hCoV-19/England/QEUH-BCCDFF/2020\|EPI_ISL_704560\|20 | ···················· | ···················· | ···················· | ···················· | ···················· | ···················· |
| W | 7791 | hCoV-19/England/QEUH-BCDDD0/2020\|EPI_ISL_704561\|20 | ···················· | ···················· | ···················· | ···················· | ···················· | ···················· |
| W | 7792 | hCoV-19/England/QEUH-BCBB05/2020\|EPI_ISL_704565\|20 | ···················· | ···················· | ···················· | ···················· | ···················· | ···················· |
| W | 7793 | hCoV-19/England/QEUH-BCB507/2020\|EPI_ISL_704566\|20 | ···················· | ···················· | ···················· | ···················· | ···················· | ···················· |
| W | 7794 | hCoV-19/England/QEUH-BCE4E9/2020\|EPI_ISL_704568\|20 | ···················· | ···················· | ···················· | ···················· | ···················· | ···················· |
| W | 7795 | hCoV-19/England/QEUH-BCBB32/2020\|EPI_ISL_704569\|20 | ···················· | ···················· | ···················· | ···················· | ···················· | ···················· |
| W | 7796 | hCoV-19/England/QEUH-BCE8B0/2020\|EPI_ISL_704571\|20 | ···················· | ···················· | ···················· | ···················· | ···················· | ···················· |
| W | 7797 | hCoV-19/England/LIVE-DC2BB1/2020\|EPI_ISL_705839\|20 | ···················· | ···················· | ···················· | ···················· | ···················· | ···················· |
| W | 7798 | hCoV-19/England/LIVE-DC2C09/2020\|EPI_ISL_705840\|20 | ···················· | ···················· | ···················· | ···················· | ···················· | ···················· |
| W | 7799 | hCoV-19/England/LIVE-DC2C63/2020\|EPI_ISL_705841\|20 | ···················· | ···················· | ···················· | ···················· | ···················· | ···················· |
| W | 7800 | hCoV-19/England/LIVE-DC2C81/2020\|EPI_ISL_705842\|20 | ···················· | ···················· | ···················· | ···················· | ···················· | ···················· |
| W | 7801 | hCoV-19/England/LIVE-DC2CDC/2020\|EPI_ISL_705843\|20 | ···················· | ···················· | ···················· | ···················· | ···················· | ···················· |
| W | 7802 | hCoV-19/England/LIVE-DC2CEB/2020\|EPI_ISL_705844\|20 | ···················· | ···················· | ···················· | ···················· | ···················· | ···················· |
| W | 7803 | hCoV-19/England/LIVE-DC2CFA/2020\|EPI_ISL_705845\|20 | ···················· | ···················· | ···················· | ···················· | ···················· | ···················· |
| W | 7804 | hCoV-19/England/LIVE-DC2D8E/2020\|EPI_ISL_705846\|20 | ···················· | ···················· | ···················· | ···················· | ···················· | ···················· |
| W | 7805 | hCoV-19/England/LIVE-DC2DD9/2020\|EPI_ISL_705847\|20 | ···················· | ···················· | ···················· | ···················· | ···················· | ···················· |
| W | 7806 | hCoV-19/England/LIVE-DC2E03/2020\|EPI_ISL_705848\|20 | ···················· | ···················· | ···················· | ···················· | ···················· | ···················· |
| W | 7807 | hCoV-19/England/LIVE-DC2E4F/2020\|EPI_ISL_705849\|20 | ···················· | ···················· | ···················· | ···················· | ···················· | ···················· |
| W | 7808 | hCoV-19/England/LIVE-DC2E8B/2020\|EPI_ISL_705850\|20 | ···················· | ···················· | ···················· | ···················· | ···················· | ···················· |
| W | 7809 | hCoV-19/England/LIVE-DC2E9A/2020\|EPI_ISL_705851\|20 | ···················· | ···················· | ···················· | ···················· | ···················· | ···················· |
| W | 7810 | hCoV-19/England/LIVE-DC2EC7/2020\|EPI_ISL_705852\|20 | ···················· | ···················· | ···················· | ···················· | ···················· | ···················· |
| W | 7811 | hCoV-19/England/LIVE-DC2F1F/2020\|EPI_ISL_705853\|20 | ···················· | ···················· | ···················· | ···················· | ···················· | ···················· |
| W | 7812 | hCoV-19/England/LIVE-DC2F4C/2020\|EPI_ISL_705854\|20 | ···················· | ···················· | ···················· | ···················· | ···················· | ···················· |
| W | 7813 | hCoV-19/England/LIVE-DC2F5B/2020\|EPI_ISL_705855\|20 | ···················· | ···················· | ···················· | ···················· | ···················· | ···················· |
| W | 7814 | hCoV-19/England/LIVE-DC2FC4/2020\|EPI_ISL_705856\|20 | ···················· | ···················· | ···················· | ···················· | ···················· | ···················· |
| W | 7815 | hCoV-19/England/LIVE-DC2FD3/2020\|EPI_ISL_705857\|20 | ···················· | ···················· | ···················· | ···················· | ···················· | ···················· |
| W | 7816 | hCoV-19/England/LIVE-DC3057/2020\|EPI_ISL_705858\|20 | ···················· | ···················· | ···················· | ···················· | ···················· | ···················· |
| W | 7817 | hCoV-19/England/LIVE-DC3093/2020\|EPI_ISL_705859\|20 | ···················· | ···················· | ···················· | ···················· | ···················· | ···················· |
| W | 7818 | hCoV-19/England/LIVE-DC3B1A/2020\|EPI_ISL_705869\|20 | ···················· | ········T··········· | ···················· | ···················· | ···················· | ···················· |
| W | 7819 | hCoV-19/England/LIVE-DC3B38/2020\|EPI_ISL_705870\|20 | ···················· | ···················· | ···················· | ···················· | ···················· | ···················· |
| W | 7820 | hCoV-19/England/LIVE-DC3BCF/2020\|EPI_ISL_705871\|20 | ···················· | ········T··········· | ···················· | ···················· | ···················· | ···················· |
| W | 7825 | hCoV-19/England/LIVE-DC3CBD/2020\|EPI_ISL_705876\|20 | ···················· | ···················· | ···················· | ···················· | ···················· | ···················· |
| W | 7827 | hCoV-19/England/LIVE-DC3D6F/2020\|EPI_ISL_705878\|20 | ···················· | ···················· | ···················· | ···················· | ···················· | ···················· |
| W | 7828 | hCoV-19/England/LIVE-DC3DF6/2020\|EPI_ISL_705879\|20 | ···················· | ···················· | ···················· | ···················· | ···················· | ···················· |
| W | 7829 | hCoV-19/England/LIVE-DC3E4E/2020\|EPI_ISL_705880\|20 | ···················· | ···················· | ···················· | ···················· | ···················· | ···················· |
| W | 7830 | hCoV-19/England/LIVE-DC3EB7/2020\|EPI_ISL_705881\|20 | ···················· | ···················· | ···················· | ···················· | ···················· | ···················· |
| W | 7831 | hCoV-19/England/LIVE-DC3F87/2020\|EPI_ISL_705882\|20 | ···················· | ·A·················· | ···················· | ···················· | ···················· | ··A················· |
| W | 7832 | hCoV-19/England/LIVE-DC3FA5/2020\|EPI_ISL_705883\|20 | ···················· | ···················· | ···················· | ···················· | ···················· | ···················· |
| W | 7833 | hCoV-19/England/LIVE-DC40A1/2020\|EPI_ISL_705884\|20 | ···················· | ···················· | ···················· | ···················· | ···················· | ···················· |
| W | 7834 | hCoV-19/England/LIVE-DC40ED/2020\|EPI_ISL_705885\|20 | ···················· | ···················· | ···················· | ···················· | ···················· | ···················· |
| W | 7835 | hCoV-19/England/LIVE-DC4126/2020\|EPI_ISL_705886\|20 | ···················· | ···················· | ···················· | ···················· | ···················· | ···················· |
| W | 7836 | hCoV-19/England/LIVE-DC4180/2020\|EPI_ISL_705887\|20 | ···················· | ···················· | ···················· | ···················· | ···················· | ···················· |
| W | 7837 | hCoV-19/England/LIVE-DC41BD/2020\|EPI_ISL_705888\|20 | ···················· | ···················· | ···················· | ···················· | ···················· | ···················· |
| W | 7838 | hCoV-19/England/LIVE-DC41EA/2020\|EPI_ISL_705889\|20 | ···················· | ···················· | ···················· | ···················· | ···················· | ···················· |
| W | 7839 | hCoV-19/England/LIVE-DC41F9/2020\|EPI_ISL_705890\|20 | ···················· | ···················· | ···················· | ···················· | ···················· | ··A················· |
| W | 7840 | hCoV-19/England/LIVE-DC42F6/2020\|EPI_ISL_705891\|20 | ···················· | ···················· | ···················· | ···················· | ···················· | ···················· |
| W | 7841 | hCoV-19/England/LIVE-DC4469/2020\|EPI_ISL_705892\|20 | ···················· | ···················· | ···················· | ···················· | ···················· | ···················· |
| W | 7842 | hCoV-19/England/LIVE-DC4584/2020\|EPI_ISL_705894\|20 | ···················· | ········T··········· | ···················· | ···················· | ···················· | ···················· |
| W | 7843 | hCoV-19/England/LIVE-DC45A2/2020\|EPI_ISL_705895\|20 | ···················· | ···················· | ···················· | ···················· | ···················· | ···················· |
| W | 7844 | hCoV-19/England/LIVE-DC45B1/2020\|EPI_ISL_705896\|20 | ···················· | ···················· | ···················· | ···················· | ···················· | ···················· |
| W | 7845 | hCoV-19/England/LIVE-DC4609/2020\|EPI_ISL_705897\|20 | ···················· | ···················· | ···················· | ···················· | ···················· | ··A················· |
| W | 7846 | hCoV-19/England/LIVE-DC4636/2020\|EPI_ISL_705898\|20 | ···················· | ···················· | ···················· | ···················· | ···················· | ···················· |
| W | 7847 | hCoV-19/England/LIVE-DC4654/2020\|EPI_ISL_705899\|20 | ···················· | ···················· | ···················· | ···················· | ···················· | ···················· |
| X | 1 | hCoV-19/Denmark/DCGC-16502/2020\|EPI_ISL_714804\|202 | ···················· | ···················· | ···················· | ···················· | ···················· | ···················· |

| | | | | | | | | |
|---|---|---|---|---|---|---|---|---|
| X | 2 | hCoV-19/Denmark/DCGC-16503/2020\|EPI_ISL_714805\|202 | ···················· | ···················· | ····················· | ···················· | ···················· | ···················· |
| X | 3 | hCoV-19/Denmark/DCGC-16504/2020\|EPI_ISL_714806\|202 | ···················· | ···················· | ····················· | ···················· | ···················· | ···················· |
| X | 4 | hCoV-19/Denmark/DCGC-16507/2020\|EPI_ISL_714807\|202 | ···················· | ···················· | ····················· | ···················· | ···················· | ···················· |
| X | 5 | hCoV-19/Denmark/DCGC-16511/2020\|EPI_ISL_714808\|202 | ···················· | ···················· | ····················· | ···················· | ···················· | ···················· |
| X | 6 | hCoV-19/Denmark/DCGC-16515/2020\|EPI_ISL_714809\|202 | ···················· | ·A·················· | ····················· | ···················· | ···················· | ···················· |
| X | 7 | hCoV-19/Denmark/DCGC-16517/2020\|EPI_ISL_714810\|202 | ···················· | ···················· | ····················· | ···················· | ···················· | ···················· |
| X | 8 | hCoV-19/Denmark/DCGC-16518/2020\|EPI_ISL_714811\|202 | ···················· | ···················· | ····················· | ···················· | ···················· | ···················· |
| X | 9 | hCoV-19/Denmark/DCGC-16520/2020\|EPI_ISL_714812\|202 | ···················· | ···················· | ····················· | ···················· | ···················· | ···················· |
| X | 10 | hCoV-19/Denmark/DCGC-16524/2020\|EPI_ISL_714813\|202 | ···················· | ···················· | ····················· | ···················· | ···················· | ···················· |
| X | 11 | hCoV-19/Denmark/DCGC-16525/2020\|EPI_ISL_714814\|202 | ···················· | ···················· | ····················· | ···················· | ···················· | ···················· |
| X | 12 | hCoV-19/Denmark/DCGC-16531/2020\|EPI_ISL_714819\|202 | ···················· | ···················· | ····················· | ···················· | ···················· | ···················· |
| X | 13 | hCoV-19/Malaysia/IMR-CV140273/2020\|EPI_ISL_718301\| | ···················· | ·A·················· | ····················· | ···················· | ···················· | ···················· |
| X | 14 | hCoV-19/Malaysia/IMR-CV140277/2020\|EPI_ISL_718302\| | ···················· | ·A·················· | ····················· | ···················· | ···················· | ···················· |
| X | 15 | hCoV-19/Malaysia/IMR-CV140278/2020\|EPI_ISL_718303\| | ···················· | ·A·················· | ····················· | ···················· | ···················· | ···················· |
| X | 16 | hCoV-19/Malaysia/IMR-CV140314/2020\|EPI_ISL_718304\| | ···················· | ·A·················· | ····················· | ···················· | ···················· | ···················· |
| X | 17 | hCoV-19/Malaysia/IMR-CV140338/2020\|EPI_ISL_718305\| | ···················· | ·A·················· | ····················· | ···················· | ···················· | ···················· |
| X | 18 | hCoV-19/Malaysia/IMR-WI194/2020\|EPI_ISL_718307\|202 | ···················· | ·A·················· | ····················· | ···················· | ···················· | ···A················ |
| X | 19 | hCoV-19/Malaysia/IMR-WI95I/2020\|EPI_ISL_718308\|202 | ···················· | ·A·················· | ····················· | ···················· | ···················· | ···A················ |
| X | 20 | hCoV-19/Malaysia/IMR-WI200/2020\|EPI_ISL_718311\|202 | ···················· | ·A·················· | ····················· | ···················· | ···················· | ···················· |
| X | 21 | hCoV-19/Malaysia/IMR-WI212/2020\|EPI_ISL_718312\|202 | ···················· | ·A·················· | ····················· | ···················· | ···················· | ···················· |
| X | 22 | hCoV-19/Malaysia/IMR-WI213/2020\|EPI_ISL_718313\|202 | ···················· | ·A·················· | ····················· | ···················· | ···················· | ···················· |
| X | 23 | hCoV-19/Malaysia/IMR-WI310/2020\|EPI_ISL_718314\|202 | ···················· | ·A·················· | ····················· | ···················· | ···················· | ···A················ |
| X | 24 | hCoV-19/Malaysia/IMR-WI313/2020\|EPI_ISL_718315\|202 | ···················· | ·A·················· | ····················· | ···················· | ···················· | ···················· |
| X | 25 | hCoV-19/England/CAMC-C46688/2020\|EPI_ISL_718331\|20 | ···················· | ···················· | ····················· | ···················· | ···················· | ···················· |
| X | 26 | hCoV-19/England/CAMC-C45364/2020\|EPI_ISL_718332\|20 | ···················· | ···················· | ····················· | ···················· | ···················· | ···················· |
| X | 27 | hCoV-19/England/CAMC-C44BE5/2020\|EPI_ISL_718334\|20 | ···················· | ···················· | ····················· | ···················· | ···················· | ···················· |
| X | 28 | hCoV-19/England/CAMC-C4605D/2020\|EPI_ISL_718338\|20 | ···················· | ·A·················· | ····················· | ···················· | ···················· | ···················· |
| X | 29 | hCoV-19/England/CAMC-C46C95/2020\|EPI_ISL_718339\|20 | ···················· | ···················· | ····················· | ···················· | ······T············· | ···················· |
| X | 30 | hCoV-19/England/CAMC-C46B6B/2020\|EPI_ISL_718344\|20 | ···················· | ···················· | ····················· | ···················· | ···················· | ···················· |
| X | 31 | hCoV-19/England/CAMC-C45FE8/2020\|EPI_ISL_718351\|20 | ···················· | ···················· | ····················· | ···················· | ···················· | ···················· |
| X | 32 | hCoV-19/England/CAMC-C44D94/2020\|EPI_ISL_718352\|20 | ···················· | ···················· | ····················· | ···················· | ···················· | ···················· |
| X | 33 | hCoV-19/England/CAMC-C466F1/2020\|EPI_ISL_718357\|20 | ···················· | ···················· | ····················· | ···················· | ···················· | ···················· |
| X | 34 | hCoV-19/England/CAMC-C460B7/2020\|EPI_ISL_718358\|20 | ···················· | ···················· | ····················· | ···················· | ···················· | ···················· |
| X | 35 | hCoV-19/England/CAMC-C4665B/2020\|EPI_ISL_718360\|20 | ···················· | ···················· | ····················· | ···················· | ···················· | ···················· |
| X | 36 | hCoV-19/England/CAMC-C404EE/2020\|EPI_ISL_718361\|20 | ···················· | ···················· | ····················· | ···················· | ···················· | ···················· |
| X | 37 | hCoV-19/England/CAMC-C459AE/2020\|EPI_ISL_718365\|20 | ···················· | ···················· | ····················· | ···················· | ···················· | ···················· |
| X | 38 | hCoV-19/England/CAMC-C4606C/2020\|EPI_ISL_718367\|20 | ···················· | ···················· | ····················· | ···················· | ···················· | ···················· |
| X | 39 | hCoV-19/England/CAMC-C45EEB/2020\|EPI_ISL_718368\|20 | ···················· | ···················· | ····················· | ···················· | ······T············· | ···················· |
| X | 40 | hCoV-19/England/CAMC-C45FAC/2020\|EPI_ISL_718376\|20 | ···················· | ···················· | ····················· | ···················· | ···················· | ···················· |
| X | 41 | hCoV-19/England/CAMC-C46002/2020\|EPI_ISL_718378\|20 | ···················· | ···················· | ····················· | ···················· | ···················· | ···················· |
| X | 42 | hCoV-19/England/CAMC-C46891/2020\|EPI_ISL_718379\|20 | ···················· | ···················· | ····················· | ···················· | ···················· | ···················· |
| X | 43 | hCoV-19/England/CAMC-C452A3/2020\|EPI_ISL_718382\|20 | ···················· | ···················· | ····················· | ···················· | ···················· | ···················· |
| X | 44 | hCoV-19/England/CAMC-C46381/2020\|EPI_ISL_718385\|20 | ···················· | ···················· | ····················· | ···················· | ···················· | ···················· |
| X | 45 | hCoV-19/England/CAMC-C44B9A/2020\|EPI_ISL_718386\|20 | ···········T········ | ···················· | ····················· | ···················· | ···················· | ···················· |
| X | 46 | hCoV-19/England/CAMC-C45B5D/2020\|EPI_ISL_718389\|20 | ···················· | ···················· | ····················· | ···················· | ···················· | ···················· |
| X | 47 | hCoV-19/England/CAMC-C45531/2020\|EPI_ISL_719600\|20 | ···················· | ···················· | ····················· | ···················· | ···················· | ···················· |
| X | 48 | hCoV-19/England/CAMC-C3F37C/2020\|EPI_ISL_719602\|20 | ···················· | ···················· | ····················· | ···················· | ···················· | ···················· |
| X | 49 | hCoV-19/England/CAMC-C4323C/2020\|EPI_ISL_719604\|20 | ···················· | ···················· | ····················· | ···················· | ···················· | ···················· |
| X | 50 | hCoV-19/England/QEUH-B9EEF3/2020\|EPI_ISL_719605\|20 | ···················· | ···················· | ····················· | ···················· | ···················· | ···················· |
| X | 51 | hCoV-19/England/QEUH-BA107F/2020\|EPI_ISL_719606\|20 | ···················· | ···················· | ····················· | ···················· | ···················· | ···················· |
| X | 52 | hCoV-19/England/QEUH-B9FFC2/2020\|EPI_ISL_719607\|20 | ···················· | ···················· | ····················· | ···················· | ···················· | ···················· |
| X | 53 | hCoV-19/England/QEUH-BA16C8/2020\|EPI_ISL_719608\|20 | ···················· | ···················· | ····················· | ···················· | ···················· | ···················· |
| X | 54 | hCoV-19/England/QEUH-BA008F/2020\|EPI_ISL_719609\|20 | ···················· | ···················· | ····················· | ···················· | ···················· | ···················· |
| X | 55 | hCoV-19/England/QEUH-BA16E6/2020\|EPI_ISL_719610\|20 | ···················· | ···················· | ····················· | ···················· | ···················· | ···················· |
| X | 56 | hCoV-19/England/QEUH-BA167D/2020\|EPI_ISL_719611\|20 | ···················· | ···················· | ····················· | ···················· | ···················· | ···················· |
| X | 57 | hCoV-19/England/QEUH-BA073F/2020\|EPI_ISL_719612\|20 | ···················· | ·A·················· | ····················· | ···················· | ···················· | ···A················ |
| X | 58 | hCoV-19/England/QEUH-BA0D1F/2020\|EPI_ISL_719613\|20 | ···················· | ···················· | ····················· | ···················· | ···················· | ···················· |
| X | 59 | hCoV-19/England/QEUH-BA166E/2020\|EPI_ISL_719617\|20 | ···················· | ···················· | ····················· | ···················· | ···················· | ···················· |
| X | 60 | hCoV-19/England/QEUH-B9EC08/2020\|EPI_ISL_719618\|20 | ···················· | ···················· | ····················· | ···················· | ···················· | ···················· |
| X | 61 | hCoV-19/England/QEUH-B9E594/2020\|EPI_ISL_719619\|20 | ···················· | ···················· | ····················· | ···················· | ···················· | ···················· |
| X | 62 | hCoV-19/England/QEUH-BA015F/2020\|EPI_ISL_719620\|20 | ···················· | ···················· | ····················· | ···················· | ···················· | ···A················ |
| X | 63 | hCoV-19/England/QEUH-BA016E/2020\|EPI_ISL_719621\|20 | ···················· | ···················· | ····················· | ···················· | ···················· | ···················· |
| X | 64 | hCoV-19/England/QEUH-BA0DC4/2020\|EPI_ISL_719622\|20 | ···················· | ···················· | ····················· | ···················· | ···················· | ···················· |
| X | 65 | hCoV-19/England/QEUH-BA076C/2020\|EPI_ISL_719623\|20 | ···················· | ···················· | ····················· | ···················· | ···················· | ···················· |
| X | 66 | hCoV-19/England/QEUH-BA1A35/2020\|EPI_ISL_719624\|20 | ···················· | ···················· | ····················· | ···················· | ···················· | ···················· |
| X | 67 | hCoV-19/England/QEUH-BA0377/2020\|EPI_ISL_719625\|20 | ···················· | ···················· | ····················· | ···················· | ···················· | ···················· |
| X | 68 | hCoV-19/England/QEUH-B9EF96/2020\|EPI_ISL_719626\|20 | ···················· | ···················· | ····················· | ···················· | ···················· | ···················· |
| X | 69 | hCoV-19/England/QEUH-BA1789/2020\|EPI_ISL_719629\|20 | ···················· | ···················· | ····················· | ···················· | ···················· | ···················· |
| X | 70 | hCoV-19/England/QEUH-BA173E/2020\|EPI_ISL_719630\|20 | ···················· | ······R············· | ····················· | ···················· | ···················· | ···················· |
| X | 71 | hCoV-19/England/QEUH-BA0D6A/2020\|EPI_ISL_719631\|20 | ···················· | ···················· | ····················· | ···················· | ···················· | ···················· |
| X | 72 | hCoV-19/England/QEUH-BA0702/2020\|EPI_ISL_719633\|20 | ···················· | ···················· | ····················· | ···················· | ······T············· | ···················· |
| X | 73 | hCoV-19/England/QEUH-BA066F/2020\|EPI_ISL_719634\|20 | ···················· | ·A·················· | ····················· | ···················· | ···················· | ···A················ |
| X | 74 | hCoV-19/England/QEUH-B9EF3C/2020\|EPI_ISL_719635\|20 | ···················· | ···················· | ····················· | ···················· | ···················· | ···················· |
| X | 75 | hCoV-19/England/QEUH-BA0535/2020\|EPI_ISL_719636\|20 | ···················· | ···················· | ····················· | ···················· | ···················· | ···················· |
| X | 76 | hCoV-19/England/QEUH-B9EFB4/2020\|EPI_ISL_719640\|20 | ···················· | ···················· | ····················· | ···················· | ···················· | ···················· |
| X | 77 | hCoV-19/England/QEUH-BA0AAF/2020\|EPI_ISL_719641\|20 | ···················· | ···················· | ····················· | ···················· | ···················· | ···················· |
| X | 79 | hCoV-19/England/QEUH-BA077B/2020\|EPI_ISL_719643\|20 | ···················· | ···················· | ····················· | ···················· | ···················· | ···················· |
| X | 80 | hCoV-19/England/QEUH-BA0492/2020\|EPI_ISL_719644\|20 | ···················· | ·A·················· | ····················· | ···················· | ···················· | ···················· |
| X | 81 | hCoV-19/England/QEUH-B9EFA5/2020\|EPI_ISL_719646\|20 | ···················· | ···················· | ····················· | ···················· | ···················· | ···················· |
| X | 82 | hCoV-19/England/QEUH-BA06E7/2020\|EPI_ISL_719648\|20 | ···················· | ···················· | ····················· | ···················· | ···················· | ···················· |

| | | | | | | | |
|---|---|---|---|---|---|---|---|
| X | 83 hCoV-19/England/QEUH-BA1956/2020\|EPI_ISL_719650\|20 | ···················· | ···················· | ···················· | ···················· | ···················· | ···················· |
| X | 84 hCoV-19/England/QEUH-BA0553/2020\|EPI_ISL_719651\|20 | ···················· | ···················· | ···················· | ···················· | ···················· | ···················· |
| X | 85 hCoV-19/England/QEUH-BA1A44/2020\|EPI_ISL_719652\|20 | ···················· | ···················· | ···················· | ···················· | ···················· | ···················· |
| X | 86 hCoV-19/England/QEUH-BA0D3D/2020\|EPI_ISL_719653\|20 | ···················· | ···················· | ···················· | ···················· | ···················· | ···················· |
| X | 87 hCoV-19/England/QEUH-BA0939/2020\|EPI_ISL_719655\|20 | ···················· | ···················· | ···················· | ···················· | ···················· | ···················· |
| X | 88 hCoV-19/England/QEUH-BA1A17/2020\|EPI_ISL_719656\|20 | ···················· | ···················· | ···················· | ···················· | ···················· | ···················· |
| X | 89 hCoV-19/England/QEUH-BA0FEC/2020\|EPI_ISL_719657\|20 | ···················· | ···················· | ···················· | ···················· | ···················· | ···················· |
| X | 90 hCoV-19/England/QEUH-BA04B0/2020\|EPI_ISL_719658\|20 | ···················· | ···················· | ···················· | ···················· | ···················· | ···················· |
| X | 91 hCoV-19/England/QEUH-B9E549/2020\|EPI_ISL_719660\|20 | ···················· | ···················· | ···················· | ···················· | ···················· | ···················· |
| X | 92 hCoV-19/England/QEUH-BA1710/2020\|EPI_ISL_719661\|20 | ···················· | ···················· | ···················· | ···················· | ···················· | ···················· |
| X | 93 hCoV-19/England/QEUH-B9EEE4/2020\|EPI_ISL_719663\|20 | ···················· | ···················· | ···················· | ···················· | ···················· | ···················· |
| X | 94 hCoV-19/England/QEUH-BA0966/2020\|EPI_ISL_719664\|20 | ···················· | ···················· | ···················· | ···················· | ···················· | ···················· |
| X | 95 hCoV-19/England/QEUH-B9EBB0/2020\|EPI_ISL_719665\|20 | ···················· | ···················· | ···················· | ···················· | ···················· | ···················· |
| X | 96 hCoV-19/England/QEUH-BA0D97/2020\|EPI_ISL_719666\|20 | ···················· | ·A·················· | ···················· | ···················· | ···················· | ···················· |
| X | 97 hCoV-19/England/QEUH-B9E479/2020\|EPI_ISL_719668\|20 | ···················· | ···················· | ···················· | ···················· | ···················· | ···················· |
| X | 98 hCoV-19/England/QEUH-BA0438/2020\|EPI_ISL_719669\|20 | ···················· | ·A··G··············· | ···················· | ···················· | ···················· | ···················· |
| X | 99 hCoV-19/England/QEUH-BA0B06/2020\|EPI_ISL_719670\|20 | ···················· | ···················· | ···················· | ···················· | ···················· | ···················· |
| X | 100 hCoV-19/England/QEUH-B9EF4B/2020\|EPI_ISL_719671\|20 | ···················· | ···················· | ···················· | ···················· | ···················· | ···················· |
| X | 101 hCoV-19/England/QEUH-BA09C0/2020\|EPI_ISL_719672\|20 | ···················· | ···················· | ···················· | ···················· | ···················· | ···················· |
| X | 102 hCoV-19/England/QEUH-BA0526/2020\|EPI_ISL_719673\|20 | ···················· | ·A·················· | ···················· | ···················· | ···················· | ··A················· |
| X | 104 hCoV-19/England/QEUH-B9EF2D/2020\|EPI_ISL_719678\|20 | ···················· | ···················· | ···················· | ···················· | ···················· | ···················· |
| X | 105 hCoV-19/England/QEUH-B9E585/2020\|EPI_ISL_719679\|20 | ···················· | ···················· | ···················· | ···················· | ···················· | ···················· |
| X | 106 hCoV-19/England/QEUH-B9EF0F/2020\|EPI_ISL_719680\|20 | ···················· | ···················· | ···················· | ···················· | ···················· | ···················· |
| X | 107 hCoV-19/England/QEUH-BA018C/2020\|EPI_ISL_719681\|20 | ···················· | ···················· | ···················· | ···················· | ···················· | ···················· |
| X | 108 hCoV-19/England/QEUH-BA0395/2020\|EPI_ISL_719683\|20 | ···················· | ···················· | ···················· | ···················· | ···················· | ···················· |
| X | 109 hCoV-19/England/QEUH-BA169B/2020\|EPI_ISL_719684\|20 | ···················· | ···················· | ···················· | ···················· | ···················· | ···················· |
| X | 110 hCoV-19/England/QEUH-B9FEE3/2020\|EPI_ISL_719685\|20 | ···················· | ···················· | ···················· | ···················· | ···················· | ···················· |
| X | 111 hCoV-19/England/QEUH-BA0632/2020\|EPI_ISL_719686\|20 | ···················· | ···················· | ···················· | ···················· | ···················· | ···················· |
| X | 112 hCoV-19/England/QEUH-BA06BA/2020\|EPI_ISL_719687\|20 | ···················· | ···················· | ···················· | ···················· | ···················· | ···················· |
| X | 113 hCoV-19/England/QEUH-BA0122/2020\|EPI_ISL_719688\|20 | ···················· | ···················· | ···················· | ···················· | ···················· | ···················· |
| X | 114 hCoV-19/England/QEUH-BA0061/2020\|EPI_ISL_719691\|20 | ···················· | ···················· | ···················· | ···················· | ···················· | ···················· |
| X | 115 hCoV-19/England/QEUH-BA0DE2/2020\|EPI_ISL_719692\|20 | ···················· | ···················· | ···················· | ···················· | ···················· | ···················· |
| X | 116 hCoV-19/England/QEUH-BA00DA/2020\|EPI_ISL_719693\|20 | ···················· | ···················· | ···················· | ···················· | ···················· | ···················· |
| X | 117 hCoV-19/England/QEUH-BA177A/2020\|EPI_ISL_719694\|20 | ···················· | ···················· | ···················· | ···················· | ···················· | ···················· |
| X | 118 hCoV-19/England/QEUH-BA0711/2020\|EPI_ISL_719698\|20 | ···················· | ···················· | ···················· | ···················· | ···················· | ···················· |
| X | 119 hCoV-19/England/QEUH-BA0FDD/2020\|EPI_ISL_719699\|20 | ···················· | ···················· | ···················· | ···················· | ···················· | ···················· |
| X | 120 hCoV-19/England/MILK-BAC89A/2020\|EPI_ISL_720600\|20 | ···················· | ···················· | ···················· | ···················· | ···················· | ···················· |
| X | 121 hCoV-19/England/MILK-BB0839/2020\|EPI_ISL_720603\|20 | ···················· | ···················· | ···················· | ···················· | ···················· | ···················· |
| X | 122 hCoV-19/England/MILK-BB0875/2020\|EPI_ISL_720604\|20 | ···················· | ···················· | ···················· | ···················· | ···················· | ···················· |
| X | 123 hCoV-19/England/MILK-BAFD10/2020\|EPI_ISL_720605\|20 | ···················· | ···················· | ···················· | ···················· | ······T············· | ···················· |
| X | 124 hCoV-19/England/MILK-BB0040/2020\|EPI_ISL_720606\|20 | ···················· | ···················· | ···················· | ···················· | ···················· | ···············A···· |
| X | 125 hCoV-19/England/MILK-BAB901/2020\|EPI_ISL_720607\|20 | ···················· | ···················· | ···················· | ···················· | ······T············· | ···················· |
| X | 126 hCoV-19/England/MILK-BB10B8/2020\|EPI_ISL_720609\|20 | ···················· | ···················· | ···················· | ···················· | ···················· | ···················· |
| X | 127 hCoV-19/England/MILK-BABAE0/2020\|EPI_ISL_720610\|20 | ···················· | ···················· | ···················· | ···················· | ···················· | ···················· |
| X | 128 hCoV-19/England/MILK-BB1B8A/2020\|EPI_ISL_720611\|20 | ···················· | ···················· | ···················· | ···················· | ···················· | ···················· |
| X | 129 hCoV-19/England/MILK-BB211E/2020\|EPI_ISL_720615\|20 | ···················· | ···················· | ···················· | ···················· | ···················· | ···················· |
| X | 130 hCoV-19/England/MILK-BABA4A/2020\|EPI_ISL_720616\|20 | ···················· | ···················· | ···················· | ···················· | ···················· | ···················· |
| X | 131 hCoV-19/England/MILK-BB1B20/2020\|EPI_ISL_720618\|20 | ···················· | ···················· | ···················· | ···················· | ···················· | ···················· |
| X | 132 hCoV-19/England/MILK-BAFCF5/2020\|EPI_ISL_720619\|20 | ···················· | ···················· | ···················· | ···················· | ······T············· | ···················· |
| X | 133 hCoV-19/England/MILK-BB1A6F/2020\|EPI_ISL_720620\|20 | ···················· | ···················· | ···················· | ···················· | ···················· | ···················· |
| X | 134 hCoV-19/England/MILK-BB112E/2020\|EPI_ISL_720621\|20 | ···················· | ···················· | ···················· | ···················· | ···················· | ···················· |
| X | 135 hCoV-19/England/MILK-BB0C6A/2020\|EPI_ISL_720622\|20 | ···················· | ···················· | ···················· | ···················· | ···················· | ···················· |
| X | 136 hCoV-19/England/MILK-BB03CF/2020\|EPI_ISL_720623\|20 | ···················· | ···················· | ···················· | ···················· | ···················· | ···················· |
| X | 137 hCoV-19/England/MILK-BB1DFD/2020\|EPI_ISL_720624\|20 | ···················· | ···················· | ···················· | ···················· | ···················· | ···················· |
| X | 138 hCoV-19/England/MILK-BAC557/2020\|EPI_ISL_720625\|20 | ···················· | ···················· | ···················· | ···················· | ···················· | ···················· |
| X | 139 hCoV-19/England/MILK-BB215A/2020\|EPI_ISL_720626\|20 | ···················· | ···················· | ···················· | ···················· | ···················· | ···················· |
| X | 140 hCoV-19/England/MILK-BB0F16/2020\|EPI_ISL_720627\|20 | ···················· | ·A·················· | ···················· | ···················· | ···················· | ···················· |
| X | 141 hCoV-19/England/MILK-BB1D57/2020\|EPI_ISL_720628\|20 | ···················· | ···················· | ···················· | ···················· | ···················· | ···················· |
| X | 142 hCoV-19/England/MILK-BB0F43/2020\|EPI_ISL_720630\|20 | ···················· | ···················· | ···················· | ···················· | ···················· | ···················· |
| X | 143 hCoV-19/England/MILK-BAC85E/2020\|EPI_ISL_720632\|20 | ···················· | ···················· | ···················· | ···················· | ···················· | ···················· |
| X | 144 hCoV-19/England/MILK-BB082A/2020\|EPI_ISL_720635\|20 | ···················· | ···················· | ···················· | ···················· | ···················· | ···················· |
| X | 145 hCoV-19/England/MILK-BB1D84/2020\|EPI_ISL_720636\|20 | ···················· | ···················· | ···················· | ···················· | ···················· | ···················· |
| X | 146 hCoV-19/England/MILK-BB0C97/2020\|EPI_ISL_720637\|20 | ···················· | ···················· | ···················· | ···················· | ···················· | ···················· |
| X | 147 hCoV-19/England/MILK-BB2002/2020\|EPI_ISL_720639\|20 | ···················· | ···················· | ···················· | ···················· | ···················· | ···················· |
| X | 148 hCoV-19/England/MILK-BB21D2/2020\|EPI_ISL_720640\|20 | ···················· | ···················· | ···················· | ···················· | ···················· | ···················· |
| X | 149 hCoV-19/England/MILK-BB1B11/2020\|EPI_ISL_720642\|20 | ···················· | ···················· | ···················· | ···················· | ···················· | ···················· |
| X | 150 hCoV-19/England/MILK-BAC5C0/2020\|EPI_ISL_720643\|20 | ···················· | ···················· | ···················· | ···················· | ···················· | ···················· |
| X | 151 hCoV-19/England/MILK-BB1A7E/2020\|EPI_ISL_720644\|20 | ···················· | ···················· | ···················· | ···················· | ···················· | ···················· |
| X | 152 hCoV-19/England/MILK-BB2178/2020\|EPI_ISL_720645\|20 | ···················· | ···················· | ···················· | ···················· | ···················· | ···················· |
| X | 153 hCoV-19/England/MILK-BB0CB5/2020\|EPI_ISL_720646\|20 | ···················· | ···················· | ···················· | ···················· | ···················· | ···················· |
| X | 154 hCoV-19/England/MILK-BB080C/2020\|EPI_ISL_720648\|20 | ···················· | ···················· | ···················· | ···················· | ···················· | ···················· |
| X | 155 hCoV-19/England/MILK-BB0AE8/2020\|EPI_ISL_720650\|20 | ···················· | ···················· | ···················· | ···················· | ···················· | ···················· |
| X | 156 hCoV-19/England/MILK-BB0A42/2020\|EPI_ISL_720651\|20 | ···················· | ···················· | ···················· | ···················· | ···················· | ···················· |
| X | 157 hCoV-19/England/MILK-BAC5B1/2020\|EPI_ISL_720653\|20 | ···················· | ···················· | ···················· | ···················· | ···················· | ···················· |
| X | 158 hCoV-19/England/MILK-BAFC9B/2020\|EPI_ISL_720654\|20 | ···················· | ···················· | ···················· | ···················· | ···················· | ···················· |
| X | 159 hCoV-19/England/MILK-BAC803/2020\|EPI_ISL_720655\|20 | ···················· | ···················· | ···················· | ···················· | ···················· | ···················· |
| X | 160 hCoV-19/England/MILK-BB0DEF/2020\|EPI_ISL_720656\|20 | ···················· | ···················· | ···················· | ···················· | ···················· | ···················· |
| X | 161 hCoV-19/England/MILK-BAC2F6/2020\|EPI_ISL_720657\|20 | ···················· | ···················· | ···················· | ···················· | ···················· | ···················· |
| X | 162 hCoV-19/England/MILK-BB1A23/2020\|EPI_ISL_720658\|20 | ···················· | ···················· | ···················· | ···················· | ···················· | ···················· |
| X | 163 hCoV-19/England/MILK-BAC84F/2020\|EPI_ISL_720660\|20 | ···················· | ···················· | ···················· | ···················· | ···················· | ···················· |

| | | | | | | | | |
|---|---|---|---|---|---|---|---|---|
| X | 164 | hCoV-19/England/MILK-BB0C1F/2020\|EPI_ISL_720661\|20 | ···················· | ···················· | ····················· | ···················· | ···················· | ···················· |
| X | 165 | hCoV-19/England/MILK-BAC311/2020\|EPI_ISL_720662\|20 | ···················· | ···················· | ····················· | ···················· | ···················· | ···················· |
| X | 166 | hCoV-19/England/MILK-BABFA5/2020\|EPI_ISL_720663\|20 | ···················· | ···················· | ····················· | ···················· | ···················· | ···················· |
| X | 167 | hCoV-19/England/MILK-BAC87C/2020\|EPI_ISL_720665\|20 | ···················· | ···················· | ····················· | ···················· | ···················· | ···················· |
| X | 168 | hCoV-19/England/MILK-BB008C/2020\|EPI_ISL_720666\|20 | ···················· | ···················· | ····················· | ···················· | ···················· | ···················· |
| X | 169 | hCoV-19/England/MILK-BABF3C/2020\|EPI_ISL_720667\|20 | ···················· | ···················· | ····················· | ···················· | ···················· | ···················· |
| X | 170 | hCoV-19/England/QEUH-C01258/2020\|EPI_ISL_720668\|20 | ···················· | ···················· | ····················· | ···················· | ···················· | ···················· |
| X | 171 | hCoV-19/England/MILK-BFD618/2020\|EPI_ISL_720671\|20 | ···················· | ···················· | ····················· | ···················· | ···················· | ···················· |
| X | 172 | hCoV-19/England/QEUH-C01249/2020\|EPI_ISL_720672\|20 | ···················· | ···················· | ····················· | ···················· | ···················· | ···················· |
| X | 173 | hCoV-19/England/MILK-BFDEC5/2020\|EPI_ISL_720673\|20 | ···················· | ···················· | ····················· | ···················· | ···················· | ···················· |
| X | 174 | hCoV-19/England/MILK-BFF722/2020\|EPI_ISL_720679\|20 | ···················· | ···················· | ····················· | ···················· | ···················· | ···················· |
| X | 175 | hCoV-19/England/MILK-BFCE99/2020\|EPI_ISL_720681\|20 | ···················· | ···················· | ····················· | ···················· | ···················· | ···················· |
| X | 176 | hCoV-19/England/MILK-BFCBB0/2020\|EPI_ISL_720687\|20 | ···················· | ·A·················· | ····················· | ···················· | ···················· | ···A················ |
| X | 177 | hCoV-19/England/ALDP-C04AB7/2020\|EPI_ISL_720688\|20 | ···················· | ···················· | ····················· | ···················· | ···················· | ···················· |
| X | 178 | hCoV-19/England/MILK-BFCF0F/2020\|EPI_ISL_720691\|20 | ···················· | ···················· | ····················· | ···················· | ···················· | ···················· |
| X | 179 | hCoV-19/England/ALDP-C049D8/2020\|EPI_ISL_720694\|20 | ···················· | ···················· | ····················· | ···················· | ···················· | ···················· |
| X | 180 | hCoV-19/England/MILK-BFCBFC/2020\|EPI_ISL_720696\|20 | ···················· | ···················· | ····················· | ···················· | ···················· | ···················· |
| X | 181 | hCoV-19/England/ALDP-C0498D/2020\|EPI_ISL_720697\|20 | ···················· | ···················· | ····················· | ···················· | ···················· | ···················· |
| X | 182 | hCoV-19/England/MILK-C006D5/2020\|EPI_ISL_720698\|20 | ···················· | ···················· | ····················· | ···················· | ···················· | ···················· |
| X | 183 | hCoV-19/Switzerland/TI-ETHZ-380305/2020\|EPI_ISL_72 | ···················· | ···················· | ····················· | ···················· | ···················· | ···················· |
| X | 184 | hCoV-19/Switzerland/TI-ETHZ-380308/2020\|EPI_ISL_72 | ···················· | ···················· | ····················· | ···················· | ···················· | ···················· |
| X | 185 | hCoV-19/Switzerland/TI-ETHZ-380311/2020\|EPI_ISL_72 | ···················· | ···················· | ····················· | ···················· | ···················· | ···················· |
| X | 186 | hCoV-19/Switzerland/VD-ETHZ-340111/2020\|EPI_ISL_72 | ···················· | ···················· | ····················· | ···················· | ···················· | ···················· |
| X | 187 | hCoV-19/Switzerland/VD-ETHZ-340113/2020\|EPI_ISL_72 | ···················· | ···················· | ····················· | ···················· | ···················· | ···················· |
| X | 188 | hCoV-19/Switzerland/VD-ETHZ-380202/2020\|EPI_ISL_72 | ···················· | ···················· | ····················· | ···················· | ···················· | ···················· |
| X | 189 | hCoV-19/Switzerland/VD-ETHZ-380226/2020\|EPI_ISL_72 | ···················· | ···················· | ····················· | ···················· | ···················· | ···················· |
| X | 190 | hCoV-19/Switzerland/VD-ETHZ-390324/2020\|EPI_ISL_72 | ···················· | ···················· | ····················· | ···················· | ···················· | ···················· |
| X | 191 | hCoV-19/Switzerland/VD-ETHZ-390414/2020\|EPI_ISL_72 | ···················· | ···················· | ····················· | ···················· | ···················· | ···················· |
| X | 192 | hCoV-19/Switzerland/VS-ETHZ-340153/2020\|EPI_ISL_72 | ···················· | ···················· | ····················· | ···················· | ···················· | ···················· |
| X | 193 | hCoV-19/Switzerland/VS-ETHZ-340165/2020\|EPI_ISL_72 | ···················· | ···················· | ····················· | ···················· | ···················· | ···················· |
| X | 194 | hCoV-19/Switzerland/VS-ETHZ-340168/2020\|EPI_ISL_72 | ···················· | ···················· | ····················· | ···················· | ···················· | ···················· |
| X | 195 | hCoV-19/Switzerland/VS-ETHZ-380207/2020\|EPI_ISL_72 | ···················· | ···················· | ····················· | ···················· | ···················· | ···················· |
| X | 196 | hCoV-19/Switzerland/VS-ETHZ-380208/2020\|EPI_ISL_72 | ···················· | ···················· | ····················· | ···················· | ···················· | ···················· |
| X | 197 | hCoV-19/Switzerland/VS-ETHZ-380210/2020\|EPI_ISL_72 | ···················· | ···················· | ····················· | ···················· | ···················· | ···················· |
| X | 198 | hCoV-19/Switzerland/VS-ETHZ-380222/2020\|EPI_ISL_72 | ···················· | ···················· | ····················· | ···················· | ···················· | ···················· |
| X | 199 | hCoV-19/Switzerland/VS-ETHZ-390213/2020\|EPI_ISL_72 | ···················· | ···················· | ····················· | ···················· | ······K············· | ···················· |
| X | 200 | hCoV-19/Switzerland/VS-ETHZ-390214/2020\|EPI_ISL_72 | ···················· | ···················· | ····················· | ···················· | ···················· | ···················· |
| X | 201 | hCoV-19/Switzerland/VS-ETHZ-390398/2020\|EPI_ISL_72 | ···················· | ···················· | ····················· | ···················· | ···················· | ···················· |
| X | 202 | hCoV-19/Switzerland/ZH-ETHZ-340117/2020\|EPI_ISL_72 | ···················· | ···················· | ····················· | ···················· | ···················· | ···················· |
| X | 203 | hCoV-19/Switzerland/ZH-ETHZ-340121/2020\|EPI_ISL_72 | ···················· | ···················· | ····················· | ···················· | ···················· | ···················· |
| X | 204 | hCoV-19/Switzerland/ZH-ETHZ-340122/2020\|EPI_ISL_72 | ···················· | ···················· | ····················· | ···················· | ···················· | ···················· |
| X | 205 | hCoV-19/Switzerland/ZH-ETHZ-340124/2020\|EPI_ISL_72 | ···················· | ···················· | ····················· | ···················· | ···················· | ···················· |
| X | 206 | hCoV-19/Switzerland/ZH-ETHZ-340125/2020\|EPI_ISL_72 | ···················· | ···················· | ····················· | ···················· | ······K············· | ···················· |
| X | 207 | hCoV-19/Switzerland/ZH-ETHZ-340127/2020\|EPI_ISL_72 | ···················· | ···················· | ····················· | ···················· | ···················· | ···················· |
| X | 208 | hCoV-19/Switzerland/ZH-ETHZ-340129/2020\|EPI_ISL_72 | ···················· | ···················· | ····················· | ···················· | ······K············· | ···················· |
| X | 209 | hCoV-19/Switzerland/ZH-ETHZ-340146/2020\|EPI_ISL_72 | ···················· | ···················· | ····················· | ···················· | ···················· | ···················· |
| X | 210 | hCoV-19/Switzerland/ZH-ETHZ-340163/2020\|EPI_ISL_72 | ···················· | ···················· | ····················· | ···················· | ···················· | ···················· |
| X | 211 | hCoV-19/Switzerland/ZH-ETHZ-340167/2020\|EPI_ISL_72 | ···················· | ················M··· | ····················· | ···················· | ···················· | ···················· |
| X | 212 | hCoV-19/Switzerland/ZH-ETHZ-340171/2020\|EPI_ISL_72 | ···················· | ···················· | ····················· | ···················· | ···················· | ···················· |
| X | 213 | hCoV-19/Switzerland/ZH-ETHZ-380220/2020\|EPI_ISL_72 | ···················· | ···················· | ····················· | ···················· | ···················· | ···················· |
| X | 214 | hCoV-19/Switzerland/ZH-ETHZ-380310/2020\|EPI_ISL_72 | ···················· | ···················· | ····················· | ···················· | ···················· | ···················· |
| X | 215 | hCoV-19/Switzerland/ZH-ETHZ-380358/2020\|EPI_ISL_72 | ···················· | ···················· | ····················· | ···················· | ···················· | ···················· |
| X | 216 | hCoV-19/Switzerland/ZH-ETHZ-380359/2020\|EPI_ISL_72 | ···················· | ···················· | ····················· | ···················· | ···················· | ···················· |
| X | 217 | hCoV-19/Switzerland/ZH-ETHZ-380360/2020\|EPI_ISL_72 | ···················· | ···················· | ····················· | ···················· | ···················· | ···················· |
| X | 218 | hCoV-19/Switzerland/ZH-ETHZ-380361/2020\|EPI_ISL_72 | ···················· | ···················· | ····················· | ···················· | ···················· | ···················· |
| X | 219 | hCoV-19/Switzerland/ZH-ETHZ-380362/2020\|EPI_ISL_72 | ···················· | ···················· | ····················· | ···················· | ···················· | ···················· |
| X | 220 | hCoV-19/Switzerland/ZH-ETHZ-380363/2020\|EPI_ISL_72 | ···················· | ···················· | ····················· | ···················· | ···················· | ···················· |
| X | 221 | hCoV-19/Switzerland/ZH-ETHZ-380364/2020\|EPI_ISL_72 | ···················· | ···················· | ····················· | ···················· | ···················· | ···················· |
| X | 222 | hCoV-19/Switzerland/ZH-ETHZ-380367/2020\|EPI_ISL_72 | ···················· | ···················· | ····················· | ···················· | ···················· | ···················· |
| X | 223 | hCoV-19/Switzerland/ZH-ETHZ-380368/2020\|EPI_ISL_72 | ···················· | ···················· | ····················· | ···················· | ···················· | ···················· |
| X | 224 | hCoV-19/Switzerland/ZH-ETHZ-380369/2020\|EPI_ISL_72 | ···················· | ···················· | ····················· | ···················· | ···················· | ···················· |
| X | 225 | hCoV-19/Switzerland/ZH-ETHZ-380370/2020\|EPI_ISL_72 | ···················· | ···················· | ····················· | ···················· | ···················· | ···················· |
| X | 226 | hCoV-19/Switzerland/ZH-ETHZ-380371/2020\|EPI_ISL_72 | ···················· | ···················· | ····················· | ···················· | ···················· | ···················· |
| X | 227 | hCoV-19/Switzerland/ZH-ETHZ-380373/2020\|EPI_ISL_72 | ···················· | ···················· | ····················· | ···················· | ···················· | ···················· |
| X | 228 | hCoV-19/Switzerland/ZH-ETHZ-380379/2020\|EPI_ISL_72 | ···················· | ···················· | ····················· | ···················· | ······K············· | ···················· |
| X | 229 | hCoV-19/Switzerland/ZH-ETHZ-380380/2020\|EPI_ISL_72 | ···················· | ···················· | ····················· | ···················· | ······K············· | ···················· |
| X | 230 | hCoV-19/Switzerland/ZH-ETHZ-380381/2020\|EPI_ISL_72 | ···················· | ···················· | ····················· | ···················· | ···················· | ···················· |
| X | 231 | hCoV-19/Switzerland/ZH-ETHZ-380382/2020\|EPI_ISL_72 | ···················· | ···················· | ····················· | ···················· | ···················· | ···················· |
| X | 232 | hCoV-19/Switzerland/ZH-ETHZ-390215/2020\|EPI_ISL_72 | ···················· | ···················· | ····················· | ···················· | ···················· | ···················· |
| X | 233 | hCoV-19/Switzerland/ZH-ETHZ-390216/2020\|EPI_ISL_72 | ···················· | ···················· | ····················· | ···················· | ···················· | ···················· |
| X | 234 | hCoV-19/Switzerland/ZH-ETHZ-390218/2020\|EPI_ISL_72 | ···················· | ···················· | ····················· | ···················· | ···················· | ···················· |
| X | 235 | hCoV-19/Switzerland/ZH-ETHZ-390224/2020\|EPI_ISL_72 | ···················· | ···················· | ····················· | ···················· | ···················· | ···················· |
| X | 236 | hCoV-19/Switzerland/ZH-ETHZ-390225/2020\|EPI_ISL_72 | ···················· | ···················· | ····················· | ···················· | ···················· | ···················· |
| X | 237 | hCoV-19/Switzerland/ZH-ETHZ-390226/2020\|EPI_ISL_72 | ···················· | ···················· | ····················· | ···················· | ···················· | ···················· |
| X | 238 | hCoV-19/Switzerland/ZH-ETHZ-390318/2020\|EPI_ISL_72 | ···················· | ···················· | ····················· | ···················· | ···················· | ···················· |
| X | 239 | hCoV-19/Switzerland/ZH-ETHZ-390387/2020\|EPI_ISL_72 | ···················· | ···················· | ····················· | ···················· | ···················· | ···················· |
| X | 240 | hCoV-19/Switzerland/ZH-ETHZ-390392/2020\|EPI_ISL_72 | ···················· | ···················· | ····················· | ···················· | ···················· | ···················· |
| X | 241 | hCoV-19/Switzerland/ZH-ETHZ-390393/2020\|EPI_ISL_72 | ···················· | ···················· | ····················· | ···················· | ···················· | ···················· |
| X | 242 | hCoV-19/Switzerland/ZH-ETHZ-390395/2020\|EPI_ISL_72 | ···················· | ···················· | ····················· | ···················· | ···················· | ···················· |
| X | 243 | hCoV-19/Switzerland/ZH-ETHZ-390399/2020\|EPI_ISL_72 | ···················· | ···················· | ····················· | ···················· | ···················· | ···················· |

X 244 hCoV-19/Switzerland/ZH-ETHZ-390405/2020|EPI_ISL_72 ···················· ···················· ···················· ···················· ···················· ····················
X 245 hCoV-19/Switzerland/ZH-ETHZ-390406/2020|EPI_ISL_72 ···················· ·A·················· ···················· ···················· ···················· ···A················
X 246 hCoV-19/Switzerland/ZH-ETHZ-390412/2020|EPI_ISL_72 ···················· ···················· ···················· ···················· ···················· ····················
X 247 hCoV-19/Switzerland/ZH-ETHZ-390413/2020|EPI_ISL_72 ···················· ···················· ···················· ···················· ···················· ····················
X 248 hCoV-19/Switzerland/ZH-ETHZ-390416/2020|EPI_ISL_72 ···················· ···················· ···················· ···················· ···················· ····················
X 249 hCoV-19/Switzerland/ZH-ETHZ-390417/2020|EPI_ISL_72 ···················· ···················· ···················· ···················· ···················· ····················
X 250 hCoV-19/Switzerland/ZH-ETHZ-390418/2020|EPI_ISL_72 ···················· ···················· ···················· ···················· ···················· ····················
X 251 hCoV-19/Switzerland/ZH-ETHZ-390419/2020|EPI_ISL_72 ···················· ···················· ···················· ···················· ···················· ····················
X 252 hCoV-19/Switzerland/ZH-ETHZ-390421/2020|EPI_ISL_72 ···················· ···················· ···················· ···················· ···················· ····················
X 253 hCoV-19/England/CAMC-BBCB70/2020|EPI_ISL_724191|20 ···················· ···················· ···················· ···················· ···················· ····················
X 254 hCoV-19/England/CAMC-BBCBAD/2020|EPI_ISL_724193|20 ···················· ···················· ···················· ···················· ···················· ····················
X 255 hCoV-19/England/CAMC-BBCBCB/2020|EPI_ISL_724195|20 ···················· ···················· ···················· ···················· ···················· ····················
X 256 hCoV-19/England/CAMC-BBCBE9/2020|EPI_ISL_724197|20 ···················· ···················· ···················· ···················· ······T············· ····················
X 257 hCoV-19/Denmark/DCGC-13508/2020|EPI_ISL_712249|202 ···················· ···················· ···················· ···················· ···················· ····················
X 258 hCoV-19/Denmark/DCGC-12949/2020|EPI_ISL_712250|202 ···················· ···················· ···················· ···················· ···················· ····················
X 259 hCoV-19/Denmark/DCGC-12979/2020|EPI_ISL_712251|202 ···················· ···················· ···················· ···················· ···················· ····················
X 260 hCoV-19/Denmark/DCGC-12994/2020|EPI_ISL_712252|202 ···················· ···················· ···················· ···················· ···················· ····················
X 261 hCoV-19/Denmark/DCGC-13005/2020|EPI_ISL_712253|202 ···················· ···················· ···················· ···················· ···················· ····················
X 262 hCoV-19/Denmark/DCGC-13054/2020|EPI_ISL_712254|202 ···················· ···················· ···················· ···················· ···················· ····················
X 263 hCoV-19/Denmark/DCGC-13073/2020|EPI_ISL_712255|202 ···················· ···················· ···················· ···················· ···················· ····················
X 264 hCoV-19/Denmark/DCGC-13079/2020|EPI_ISL_712256|202 ···················· ···················· ···················· ···················· ···················· ····················
X 265 hCoV-19/Denmark/DCGC-13093/2020|EPI_ISL_712257|202 ···················· ···················· ···················· ···················· ···················· ····················
X 266 hCoV-19/Denmark/DCGC-13150/2020|EPI_ISL_712258|202 ···················· ···················· ···················· ···················· ···················· ····················
X 267 hCoV-19/Denmark/DCGC-13171/2020|EPI_ISL_712259|202 ···················· ···················· ···················· ···················· ···················· ····················
X 268 hCoV-19/Denmark/DCGC-13339/2020|EPI_ISL_712266|202 ···················· ···················· ···················· ···················· ···················· ····················
X 269 hCoV-19/Denmark/DCGC-13383/2020|EPI_ISL_712268|202 ···················· ···················· ···················· ···················· ···················· ····················
X 270 hCoV-19/Denmark/DCGC-13388/2020|EPI_ISL_712269|202 ···················· ···················· ···················· ···················· ···················· ····················
X 271 hCoV-19/Denmark/DCGC-13389/2020|EPI_ISL_712270|202 ···················· ···················· ···················· ···················· ···················· ····················
X 272 hCoV-19/Denmark/DCGC-13395/2020|EPI_ISL_712271|202 ···················· ···················· ···················· ···················· ···················· ····················
X 273 hCoV-19/Denmark/DCGC-13418/2020|EPI_ISL_712272|202 ···················· ···················· ···················· ···················· ···················· ····················
X 274 hCoV-19/Denmark/DCGC-13427/2020|EPI_ISL_712273|202 ···················· ···················· ···················· ···················· ···················· ····················
X 275 hCoV-19/Denmark/DCGC-13440/2020|EPI_ISL_712274|202 ···················· ···················· ···················· ···················· ···················· ····················
X 276 hCoV-19/Denmark/DCGC-13489/2020|EPI_ISL_712275|202 ···················· ···················· ···················· ···················· ···················· ····················
X 277 hCoV-19/Denmark/DCGC-13524/2020|EPI_ISL_712276|202 ···················· ···················· ···················· ···················· ···················· ····················
X 278 hCoV-19/Denmark/DCGC-13531/2020|EPI_ISL_712277|202 ···················· ···················· ···················· ···················· ···················· ····················
X 279 hCoV-19/Denmark/DCGC-13567/2020|EPI_ISL_712278|202 ···················· ···················· ···················· ···················· ···················· ····················
X 280 hCoV-19/Denmark/DCGC-13630/2020|EPI_ISL_712279|202 ···················· ···················· ···················· ···················· ···················· ····················
X 281 hCoV-19/Denmark/DCGC-13647/2020|EPI_ISL_712280|202 ···················· ···················· ···················· ···················· ···················· ····················
X 282 hCoV-19/Denmark/DCGC-13661/2020|EPI_ISL_712281|202 ···················· ···················· ···················· ···················· ···················· ····················
X 283 hCoV-19/Denmark/DCGC-13673/2020|EPI_ISL_712282|202 ···················· ···················· ···················· ···················· ···················· ····················
X 284 hCoV-19/Denmark/DCGC-13709/2020|EPI_ISL_712283|202 ···················· ···················· ···················· ···················· ···················· ····················
X 285 hCoV-19/Denmark/DCGC-13715/2020|EPI_ISL_712284|202 ···················· ···················· ···················· ···················· ···················· ····················
X 286 hCoV-19/Denmark/DCGC-13757/2020|EPI_ISL_712285|202 ···················· ···················· ···················· ···················· ···················· ····················
X 287 hCoV-19/Denmark/DCGC-13780/2020|EPI_ISL_712286|202 ···················· ···················· ···················· ···················· ···················· ····················
X 288 hCoV-19/Denmark/DCGC-13791/2020|EPI_ISL_712287|202 ···················· ···················· ···················· ···················· ···················· ····················
X 289 hCoV-19/Denmark/DCGC-13807/2020|EPI_ISL_712288|202 ···················· ···················· ···················· ···················· ···················· ····················
X 290 hCoV-19/Denmark/DCGC-13821/2020|EPI_ISL_712289|202 ···················· ···················· ···················· ···················· ···················· ····················
X 291 hCoV-19/Denmark/DCGC-14909/2020|EPI_ISL_713513|202 ···················· ···················· ···················· ···················· ···················· ····················
X 292 hCoV-19/Denmark/DCGC-14912/2020|EPI_ISL_713514|202 ···················· ···················· ···················· ···················· ···················· ····················
X 293 hCoV-19/Denmark/DCGC-14914/2020|EPI_ISL_713515|202 ···················· ···················· ···················· ···················· ···················· ····················
X 294 hCoV-19/Denmark/DCGC-14915/2020|EPI_ISL_713516|202 ···················· ···················· ···················· ···················· ···················· ····················
X 295 hCoV-19/Denmark/DCGC-14916/2020|EPI_ISL_713517|202 ···················· ···················· ···················· ···················· ···················· ····················
X 296 hCoV-19/Denmark/DCGC-14917/2020|EPI_ISL_713518|202 ···················· ···················· ···················· ···················· ···················· ····················
X 297 hCoV-19/Denmark/DCGC-14918/2020|EPI_ISL_713519|202 ···················· ··········A········· ···················· ···················· ···················· ····················
X 298 hCoV-19/Denmark/DCGC-14919/2020|EPI_ISL_713520|202 ···················· ···················· ···················· ···················· ···················· ····················
X 299 hCoV-19/Denmark/DCGC-14920/2020|EPI_ISL_713521|202 ···················· ···················· ···················· ···················· ···················· ····················
X 300 hCoV-19/Denmark/DCGC-14921/2020|EPI_ISL_713522|202 ···················· ···················· ···················· ···················· ···················· ····················
X 301 hCoV-19/Denmark/DCGC-14922/2020|EPI_ISL_713523|202 ···················· ···················· ···················· ···················· ···················· ····················
X 302 hCoV-19/Denmark/DCGC-14923/2020|EPI_ISL_713524|202 ···················· ···················· ···················· ···················· ···················· ····················
X 303 hCoV-19/Denmark/DCGC-14924/2020|EPI_ISL_713525|202 ···················· ···················· ···················· ···················· ···················· ····················
X 304 hCoV-19/Denmark/DCGC-14926/2020|EPI_ISL_713526|202 ···················· ···················· ···················· ···················· ···················· ····················
X 305 hCoV-19/Denmark/DCGC-14929/2020|EPI_ISL_713527|202 ···················· ···················· ···················· ···················· ···················· ····················
X 306 hCoV-19/Denmark/DCGC-14930/2020|EPI_ISL_713528|202 ···················· ···················· ···················· ···················· ···················· ····················
X 307 hCoV-19/Denmark/DCGC-14931/2020|EPI_ISL_713529|202 ···················· ·A·················· ···················· ···················· ···················· ···A················
X 308 hCoV-19/Denmark/DCGC-14932/2020|EPI_ISL_713530|202 ···················· ···················· ···················· ···················· ···················· ····················
X 309 hCoV-19/Denmark/DCGC-14934/2020|EPI_ISL_713531|202 ···················· ·A·················· ···················· ···················· ···················· ···A················
X 310 hCoV-19/Denmark/DCGC-14936/2020|EPI_ISL_713532|202 ···················· ···················· ···················· ···················· ···················· ····················
X 311 hCoV-19/Denmark/DCGC-14939/2020|EPI_ISL_713533|202 ···················· ···················· ···················· ···················· ···················· ····················
X 312 hCoV-19/Denmark/DCGC-14940/2020|EPI_ISL_713534|202 ···················· ···················· ···················· ···················· ······T············· ····················
X 313 hCoV-19/Denmark/DCGC-14941/2020|EPI_ISL_713535|202 ···················· ···················· ···················· ···················· ···················· ····················
X 314 hCoV-19/Denmark/DCGC-14943/2020|EPI_ISL_713536|202 ···················· ···················· ···················· ···················· ···················· ····················
X 315 hCoV-19/Denmark/DCGC-14944/2020|EPI_ISL_713537|202 ···················· ···················· ···················· ···················· ···················· ····················
X 316 hCoV-19/Denmark/DCGC-14946/2020|EPI_ISL_713538|202 ···················· ···················· ···················· ···················· ···················· ····················
X 317 hCoV-19/Denmark/DCGC-14947/2020|EPI_ISL_713539|202 ···················· ···················· ···················· ···················· ···················· ····················
X 318 hCoV-19/Denmark/DCGC-14949/2020|EPI_ISL_713540|202 ···················· ···················· ···················· ···················· ···················· ····················
X 319 hCoV-19/Denmark/DCGC-14950/2020|EPI_ISL_713541|202 ···················· ···················· ···················· ···················· ···················· ····················
X 320 hCoV-19/Denmark/DCGC-14951/2020|EPI_ISL_713542|202 ···················· ···················· ···················· ···················· ···················· ····················
X 321 hCoV-19/Denmark/DCGC-14952/2020|EPI_ISL_713543|202 ···················· ···················· ···················· ···················· ···················· ····················
X 322 hCoV-19/Denmark/DCGC-14953/2020|EPI_ISL_713544|202 ···················· ···················· ···················· ···················· ···················· ····················
X 323 hCoV-19/Denmark/DCGC-14954/2020|EPI_ISL_713545|202 ···················· ···················· ···················· ···················· ···················· ····················

| | | | | | | | | |
|---|---|---|---|---|---|---|---|---|
| X | 324 | hCoV-19/Denmark/DCGC-14955/2020\|EPI_ISL_713546\|202 | ···················· | ···················· | ····················· | ···················· | ···················· | ···················· |
| X | 325 | hCoV-19/Denmark/DCGC-14956/2020\|EPI_ISL_713547\|202 | ···················· | ···················· | ····················· | ···················· | ···················· | ···················· |
| X | 326 | hCoV-19/Denmark/DCGC-14959/2020\|EPI_ISL_713548\|202 | ···················· | ···················· | ····················· | ···················· | ···················· | ···················· |
| X | 327 | hCoV-19/Denmark/DCGC-14960/2020\|EPI_ISL_713549\|202 | ···················· | ···················· | ····················· | ···················· | ···················· | ···················· |
| X | 328 | hCoV-19/Denmark/DCGC-14961/2020\|EPI_ISL_713550\|202 | ···················· | ···················· | ····················· | ···················· | ···················· | ···················· |
| X | 329 | hCoV-19/Denmark/DCGC-14963/2020\|EPI_ISL_713551\|202 | ···················· | ···················· | ····················· | ···················· | ···················· | ···················· |
| X | 330 | hCoV-19/Denmark/DCGC-14965/2020\|EPI_ISL_713552\|202 | ···················· | ···················· | ····················· | ···················· | ···················· | ···················· |
| X | 331 | hCoV-19/Denmark/DCGC-14966/2020\|EPI_ISL_713553\|202 | ···················· | ···················· | ····················· | ···················· | ···················· | ···················· |
| X | 332 | hCoV-19/Denmark/DCGC-14967/2020\|EPI_ISL_713554\|202 | ···················· | ···················· | ····················· | ···················· | ···················· | ···················· |
| X | 333 | hCoV-19/Denmark/DCGC-14968/2020\|EPI_ISL_713555\|202 | ···················· | ···················· | ····················· | ···················· | ···················· | ···················· |
| X | 334 | hCoV-19/Denmark/DCGC-14971/2020\|EPI_ISL_713556\|202 | ···················· | ···················· | ····················· | ···················· | ···················· | ···················· |
| X | 335 | hCoV-19/Denmark/DCGC-14972/2020\|EPI_ISL_713557\|202 | ···················· | ···················· | ····················· | ···················· | ···················· | ···················· |
| X | 336 | hCoV-19/Denmark/DCGC-14975/2020\|EPI_ISL_713558\|202 | ···················· | ···················· | ····················· | ···················· | ···················· | ···················· |
| X | 337 | hCoV-19/Denmark/DCGC-14980/2020\|EPI_ISL_713559\|202 | ···················· | ···················· | ····················· | ···················· | ···················· | ···················· |
| X | 338 | hCoV-19/Denmark/DCGC-14982/2020\|EPI_ISL_713560\|202 | ···················· | ···················· | ····················· | ···················· | ···················· | ···················· |
| X | 339 | hCoV-19/Denmark/DCGC-14984/2020\|EPI_ISL_713561\|202 | ···················· | ···················· | ····················· | ···················· | ···················· | ···················· |
| X | 340 | hCoV-19/Denmark/DCGC-14985/2020\|EPI_ISL_713562\|202 | ···················· | ···················· | ····················· | ···················· | ···················· | ···················· |
| X | 341 | hCoV-19/Denmark/DCGC-14986/2020\|EPI_ISL_713563\|202 | ···················· | ···················· | ····················· | ···················· | ···················· | ···················· |
| X | 342 | hCoV-19/Denmark/DCGC-14987/2020\|EPI_ISL_713564\|202 | ···················· | ···················· | ····················· | ···················· | ···················· | ···················· |
| X | 343 | hCoV-19/Denmark/DCGC-14988/2020\|EPI_ISL_713565\|202 | ···················· | ···················· | ····················· | ···················· | ···················· | ···················· |
| X | 344 | hCoV-19/Denmark/DCGC-14990/2020\|EPI_ISL_713566\|202 | ···················· | ···················· | ····················· | ···················· | ···················· | ···················· |
| X | 345 | hCoV-19/Denmark/DCGC-14993/2020\|EPI_ISL_713567\|202 | ···················· | ···················· | ····················· | ···················· | ···················· | ···················· |
| X | 346 | hCoV-19/Denmark/DCGC-14994/2020\|EPI_ISL_713568\|202 | ···················· | ···················· | ····················· | ···················· | ···················· | ···················· |
| X | 347 | hCoV-19/Denmark/DCGC-14997/2020\|EPI_ISL_713569\|202 | ···················· | ···················· | ····················· | ···················· | ···················· | ···················· |
| X | 348 | hCoV-19/Denmark/DCGC-14999/2020\|EPI_ISL_713570\|202 | ···················· | ···················· | ····················· | ···················· | ···················· | ···················· |
| X | 349 | hCoV-19/Denmark/DCGC-15001/2020\|EPI_ISL_713571\|202 | ···················· | ···················· | ····················· | ···················· | ···················· | ···················· |
| X | 350 | hCoV-19/Denmark/DCGC-15003/2020\|EPI_ISL_713572\|202 | ···················· | ·N·················· | ····················· | ···················· | ···················· | ··A················· |
| X | 351 | hCoV-19/Denmark/DCGC-15005/2020\|EPI_ISL_713573\|202 | ···················· | ···················· | ····················· | ···················· | ···················· | ···················· |
| X | 352 | hCoV-19/Denmark/DCGC-15006/2020\|EPI_ISL_713574\|202 | ···················· | ···················· | ····················· | ···················· | ···················· | ···················· |
| X | 353 | hCoV-19/Denmark/DCGC-15007/2020\|EPI_ISL_713575\|202 | ···················· | ···················· | ····················· | ···················· | ···················· | ···················· |
| X | 354 | hCoV-19/Denmark/DCGC-15008/2020\|EPI_ISL_713576\|202 | ···················· | ···················· | ····················· | ···················· | ···················· | ···················· |
| X | 355 | hCoV-19/Denmark/DCGC-15009/2020\|EPI_ISL_713577\|202 | ···················· | ···················· | ····················· | ···················· | ···················· | ···················· |
| X | 356 | hCoV-19/Denmark/DCGC-15011/2020\|EPI_ISL_713578\|202 | ···················· | ···················· | ····················· | ···················· | ···················· | ···················· |
| X | 357 | hCoV-19/Denmark/DCGC-15016/2020\|EPI_ISL_713579\|202 | ···················· | ···················· | ····················· | ···················· | ···················· | ···················· |
| X | 358 | hCoV-19/Denmark/DCGC-15017/2020\|EPI_ISL_713580\|202 | ···················· | ···················· | ····················· | ···················· | ···················· | ···················· |
| X | 359 | hCoV-19/Denmark/DCGC-15018/2020\|EPI_ISL_713581\|202 | ···················· | ·············A······ | ····················· | ···················· | ···················· | ···················· |
| X | 360 | hCoV-19/Denmark/DCGC-15020/2020\|EPI_ISL_713582\|202 | ···················· | ···················· | ····················· | ···················· | ···················· | ···················· |
| X | 361 | hCoV-19/Denmark/DCGC-15021/2020\|EPI_ISL_713583\|202 | ···················· | ···················· | ····················· | ···················· | ···················· | ···················· |
| X | 362 | hCoV-19/Denmark/DCGC-15022/2020\|EPI_ISL_713584\|202 | ···················· | ·A·················· | ····················· | ···················· | ···················· | ··A················· |
| X | 363 | hCoV-19/Denmark/DCGC-15023/2020\|EPI_ISL_713585\|202 | ···················· | ···················· | ····················· | ···················· | ···················· | ···················· |
| X | 364 | hCoV-19/Denmark/DCGC-15024/2020\|EPI_ISL_713586\|202 | ···················· | ···················· | ····················· | ···················· | ···················· | ···················· |
| X | 365 | hCoV-19/Denmark/DCGC-15025/2020\|EPI_ISL_713587\|202 | ···················· | ···················· | ····················· | ···················· | ···················· | ···················· |
| X | 366 | hCoV-19/Denmark/DCGC-15026/2020\|EPI_ISL_713588\|202 | ···················· | ···················· | ····················· | ···················· | ···················· | ···················· |
| X | 367 | hCoV-19/Denmark/DCGC-15027/2020\|EPI_ISL_713589\|202 | ···················· | ···················· | ····················· | ···················· | ···················· | ···················· |
| X | 368 | hCoV-19/Denmark/DCGC-15037/2020\|EPI_ISL_713597\|202 | ···················· | ···················· | ····················· | ···················· | ···················· | ···················· |
| X | 369 | hCoV-19/Denmark/DCGC-15038/2020\|EPI_ISL_713598\|202 | ···················· | ···················· | ····················· | ···················· | ···················· | ···················· |
| X | 370 | hCoV-19/Denmark/DCGC-15039/2020\|EPI_ISL_713599\|202 | ···················· | ·A·················· | ····················· | ···················· | ···················· | ···················· |
| X | 371 | hCoV-19/Denmark/DCGC-16491/2020\|EPI_ISL_714800\|202 | ···················· | ···················· | ····················· | ···················· | ···················· | ···················· |
| X | 372 | hCoV-19/Denmark/DCGC-16495/2020\|EPI_ISL_714801\|202 | ···················· | ···················· | ····················· | ···················· | ···················· | ···················· |
| X | 373 | hCoV-19/Denmark/DCGC-16498/2020\|EPI_ISL_714802\|202 | ···················· | ···················· | ····················· | ···················· | ···················· | ···················· |
| X | 374 | hCoV-19/Denmark/DCGC-16501/2020\|EPI_ISL_714803\|202 | ···················· | ···················· | ····················· | ···················· | ···················· | ···················· |
| X | 375 | hCoV-19/Denmark/DCGC-16526/2020\|EPI_ISL_714815\|202 | ···················· | ···················· | ····················· | ···················· | ···················· | ···················· |
| X | 376 | hCoV-19/Denmark/DCGC-16527/2020\|EPI_ISL_714816\|202 | ···················· | ···················· | ····················· | ···················· | ···················· | ···················· |
| X | 377 | hCoV-19/Denmark/DCGC-16528/2020\|EPI_ISL_714817\|202 | ···················· | ···················· | ····················· | ···················· | ···················· | ···················· |
| X | 378 | hCoV-19/Denmark/DCGC-16530/2020\|EPI_ISL_714818\|202 | ···················· | ···················· | ····················· | ···················· | ······T············· | ···················· |
| X | 379 | hCoV-19/Denmark/DCGC-16532/2020\|EPI_ISL_714820\|202 | ···················· | ·A·················· | ····················· | ···················· | ···················· | ··A················· |
| X | 380 | hCoV-19/Denmark/DCGC-16533/2020\|EPI_ISL_714821\|202 | ···················· | ···················· | ····················· | ···················· | ···················· | ···················· |
| X | 381 | hCoV-19/Denmark/DCGC-16535/2020\|EPI_ISL_714822\|202 | ···················· | ···················· | ····················· | ···················· | ···················· | ···················· |
| X | 382 | hCoV-19/Denmark/DCGC-16536/2020\|EPI_ISL_714823\|202 | ···················· | ···················· | ····················· | ···················· | ···················· | ···················· |
| X | 383 | hCoV-19/Denmark/DCGC-16537/2020\|EPI_ISL_714824\|202 | ···················· | ···················· | ····················· | ···················· | ···················· | ···················· |
| X | 384 | hCoV-19/Denmark/DCGC-16540/2020\|EPI_ISL_714825\|202 | ···················· | ···················· | ····················· | ···················· | ···················· | ···················· |
| X | 385 | hCoV-19/Denmark/DCGC-16541/2020\|EPI_ISL_714826\|202 | ···················· | ·A·················· | ····················· | ···················· | ···················· | ···················· |
| X | 386 | hCoV-19/Denmark/DCGC-16542/2020\|EPI_ISL_714827\|202 | ···················· | ···················· | ····················· | ···················· | ···················· | ···················· |
| X | 387 | hCoV-19/Denmark/DCGC-16544/2020\|EPI_ISL_714828\|202 | ···················· | ···················· | ····················· | ···················· | ···················· | ···················· |
| X | 388 | hCoV-19/Denmark/DCGC-16546/2020\|EPI_ISL_714829\|202 | ···················· | ···················· | ····················· | ···················· | ···················· | ···················· |
| X | 389 | hCoV-19/Denmark/DCGC-16547/2020\|EPI_ISL_714830\|202 | ···················· | ···················· | ····················· | ···················· | ···················· | ···················· |
| X | 390 | hCoV-19/Denmark/DCGC-16548/2020\|EPI_ISL_714831\|202 | ···················· | ···················· | ····················· | ···················· | ···················· | ···················· |
| X | 391 | hCoV-19/Denmark/DCGC-16551/2020\|EPI_ISL_714832\|202 | ···················· | ···················· | ····················· | ···················· | ···················· | ···················· |
| X | 392 | hCoV-19/Denmark/DCGC-16553/2020\|EPI_ISL_714833\|202 | ···················· | ···················· | ····················· | ···················· | ···················· | ···················· |
| X | 393 | hCoV-19/Denmark/DCGC-16554/2020\|EPI_ISL_714834\|202 | ···················· | ···················· | ····················· | ···················· | ···················· | ···················· |
| X | 394 | hCoV-19/Denmark/DCGC-16557/2020\|EPI_ISL_714835\|202 | ···················· | ···················· | ····················· | ···················· | ···················· | ···················· |
| X | 395 | hCoV-19/Denmark/DCGC-16558/2020\|EPI_ISL_714836\|202 | ···················· | ···················· | ····················· | ···················· | ···················· | ···················· |
| X | 396 | hCoV-19/Denmark/DCGC-16559/2020\|EPI_ISL_714837\|202 | ···················· | ···················· | ····················· | ···················· | ···················· | ···················· |
| X | 397 | hCoV-19/Denmark/DCGC-16560/2020\|EPI_ISL_714838\|202 | ···················· | ···················· | ····················· | ···················· | ···················· | ···················· |
| X | 398 | hCoV-19/Denmark/DCGC-16563/2020\|EPI_ISL_714839\|202 | ···················· | ···················· | ····················· | ···················· | ···················· | ···················· |
| X | 399 | hCoV-19/Denmark/DCGC-16566/2020\|EPI_ISL_714840\|202 | ···················· | ···················· | ····················· | ···················· | ···················· | ···················· |
| X | 400 | hCoV-19/Denmark/DCGC-16568/2020\|EPI_ISL_714841\|202 | ···················· | ···················· | ····················· | ···················· | ···················· | ···················· |
| X | 401 | hCoV-19/Denmark/DCGC-16569/2020\|EPI_ISL_714842\|202 | ···················· | ···················· | ····················· | ···················· | ···················· | ···················· |
| X | 402 | hCoV-19/Denmark/DCGC-16570/2020\|EPI_ISL_714843\|202 | ···················· | ···················· | ····················· | ···················· | ···················· | ···················· |
| X | 403 | hCoV-19/Denmark/DCGC-16571/2020\|EPI_ISL_714844\|202 | ···················· | ···················· | ····················· | ···················· | ···················· | ···················· |

X 404 hCoV-19/Denmark/DCGC-16572/2020|EPI_ISL_714845|202 ···················· ···················· ····················· ···················· ···················· ····················
X 405 hCoV-19/Denmark/DCGC-16576/2020|EPI_ISL_714846|202 ···················· ···················· ····················· ···················· ···················· ····················
X 406 hCoV-19/Denmark/DCGC-16577/2020|EPI_ISL_714847|202 ···················· ···················· ····················· ···················· ···················· ····················
X 407 hCoV-19/Denmark/DCGC-16578/2020|EPI_ISL_714848|202 ···················· ···················· ····················· ···················· ···················· ····················
X 408 hCoV-19/Denmark/DCGC-16579/2020|EPI_ISL_714849|202 ···················· ···················· ····················· ···················· ···················· ····················
X 409 hCoV-19/Denmark/DCGC-16580/2020|EPI_ISL_714850|202 ···················· ···················· ····················· ···················· ···················· ····················
X 410 hCoV-19/Denmark/DCGC-16581/2020|EPI_ISL_714851|202 ···················· ···················· ····················· ···················· ···················· ····················
X 411 hCoV-19/Denmark/DCGC-16582/2020|EPI_ISL_714852|202 ···················· ···················· ····················· ···················· ···················· ····················
X 412 hCoV-19/Denmark/DCGC-16583/2020|EPI_ISL_714853|202 ···················· ···················· ····················· ···················· ···················· ····················
X 413 hCoV-19/Denmark/DCGC-16584/2020|EPI_ISL_714854|202 ···················· ···················· ····················· ···················· ···················· ····················
X 414 hCoV-19/Denmark/DCGC-16585/2020|EPI_ISL_714855|202 ···················· ···················· ····················· ···················· ···················· ····················
X 415 hCoV-19/Denmark/DCGC-16587/2020|EPI_ISL_714856|202 ···················· ···················· ····················· ···················· ···················· ····················
X 416 hCoV-19/Denmark/DCGC-16589/2020|EPI_ISL_714857|202 ···················· ···················· ····················· ···················· ···················· ····················
X 417 hCoV-19/Denmark/DCGC-16593/2020|EPI_ISL_714858|202 ···················· ···················· ····················· ···················· ···················· ····················
X 418 hCoV-19/Denmark/DCGC-16595/2020|EPI_ISL_714859|202 ···················· ···················· ····················· ···················· ···················· ····················
X 419 hCoV-19/Denmark/DCGC-16598/2020|EPI_ISL_714860|202 ···················· ···················· ····················· ···················· ···················· ····················
X 420 hCoV-19/Denmark/DCGC-16599/2020|EPI_ISL_714861|202 ···················· ···················· ····················· ···················· ···················· ····················
X 421 hCoV-19/Denmark/DCGC-16600/2020|EPI_ISL_714862|202 ···················· ···················· ····················· ···················· ···················· ····················
X 422 hCoV-19/Denmark/DCGC-16602/2020|EPI_ISL_714863|202 ···················· ···················· ····················· ···················· ···················· ····················
X 423 hCoV-19/Denmark/DCGC-16603/2020|EPI_ISL_714864|202 ···················· ···················· ····················· ···················· ···················· ····················
X 424 hCoV-19/Denmark/DCGC-16605/2020|EPI_ISL_714865|202 ···················· ···················· ····················· ···················· ···················· ····················
X 425 hCoV-19/Denmark/DCGC-16606/2020|EPI_ISL_714866|202 ···················· ···················· ····················· ···················· ···················· ····················
X 426 hCoV-19/Denmark/DCGC-16607/2020|EPI_ISL_714867|202 ···················· ···················· ····················· ···················· ···················· ····················
X 427 hCoV-19/Denmark/DCGC-16608/2020|EPI_ISL_714868|202 ···················· ···················· ····················· ···················· ···················· ····················
X 428 hCoV-19/Denmark/DCGC-16612/2020|EPI_ISL_714869|202 ···················· ···················· ····················· ···················· ···················· ····················
X 429 hCoV-19/Denmark/DCGC-16613/2020|EPI_ISL_714870|202 ···················· ···················· ····················· ···················· ···················· ····················
X 430 hCoV-19/Denmark/DCGC-16614/2020|EPI_ISL_714871|202 ···················· ···················· ····················· ···················· ···················· ····················
X 431 hCoV-19/Denmark/DCGC-16615/2020|EPI_ISL_714872|202 ···················· ···················· ····················· ···················· ···················· ····················
X 432 hCoV-19/Denmark/DCGC-16617/2020|EPI_ISL_714873|202 ···················· ···················· ····················· ···················· ···················· ····················
X 433 hCoV-19/Denmark/DCGC-16619/2020|EPI_ISL_714874|202 ···················· ···················· ····················· ···················· ···················· ····················
X 434 hCoV-19/Denmark/DCGC-16620/2020|EPI_ISL_714875|202 ···················· ···················· ····················· ···················· ···················· ····················
X 435 hCoV-19/Denmark/DCGC-16622/2020|EPI_ISL_714876|202 ···················· ·A·················· ····················· ···················· ···················· ····················
X 436 hCoV-19/Denmark/DCGC-16623/2020|EPI_ISL_714877|202 ···················· ···················· ····················· ···················· ···················· ····················
X 437 hCoV-19/Denmark/DCGC-16626/2020|EPI_ISL_714878|202 ···················· ···················· ····················· ···················· ···················· ····················
X 438 hCoV-19/Denmark/DCGC-16627/2020|EPI_ISL_714879|202 ···················· ···················· ····················· ···················· ···················· ····················
X 439 hCoV-19/Denmark/DCGC-16630/2020|EPI_ISL_714880|202 ···················· ···················· ····················· ···················· ···················· ····················
X 440 hCoV-19/Denmark/DCGC-16631/2020|EPI_ISL_714881|202 ···················· ···················· ····················· ···················· ···················· ····················
X 441 hCoV-19/Denmark/DCGC-16632/2020|EPI_ISL_714882|202 ···················· ···················· ····················· ···················· ···················· ····················
X 442 hCoV-19/Denmark/DCGC-16633/2020|EPI_ISL_714883|202 ···················· ···················· ····················· ···················· ···················· ····················
X 443 hCoV-19/Denmark/DCGC-16635/2020|EPI_ISL_714884|202 ···················· ···················· ····················· ···················· ···················· ····················
X 444 hCoV-19/Denmark/DCGC-16636/2020|EPI_ISL_714885|202 ···················· ···················· ····················· ···················· ···················· ····················
X 445 hCoV-19/Denmark/DCGC-16638/2020|EPI_ISL_714886|202 ···················· ···················· ····················· ···················· ···················· ····················
X 446 hCoV-19/Denmark/DCGC-16639/2020|EPI_ISL_714887|202 ···················· ···················· ····················· ···················· ···················· ····················
X 447 hCoV-19/Denmark/DCGC-16641/2020|EPI_ISL_714888|202 ···················· ···················· ····················· ···················· ···················· ····················
X 448 hCoV-19/Denmark/DCGC-16642/2020|EPI_ISL_714889|202 ···················· ···················· ····················· ···················· ···················· ····················
X 449 hCoV-19/Denmark/DCGC-16645/2020|EPI_ISL_714890|202 ···················· ···················· ····················· ···················· ···················· ····················
X 450 hCoV-19/Denmark/DCGC-16650/2020|EPI_ISL_714891|202 ···················· ···················· ····················· ···················· ···················· ····················
X 451 hCoV-19/Denmark/DCGC-16651/2020|EPI_ISL_714892|202 ···················· ···················· ····················· ···················· ···················· ····················
X 452 hCoV-19/Denmark/DCGC-16652/2020|EPI_ISL_714893|202 ···················· ···················· ····················· ···················· ···················· ····················
X 453 hCoV-19/Denmark/DCGC-16653/2020|EPI_ISL_714894|202 ···················· ···················· ····················· ···················· ···················· ····················
X 454 hCoV-19/Denmark/DCGC-16654/2020|EPI_ISL_714895|202 ···················· ···················· ····················· ···················· ···················· ····················
X 455 hCoV-19/Denmark/DCGC-16656/2020|EPI_ISL_714896|202 ···················· ···················· ····················· ···················· ···················· ····················
X 456 hCoV-19/Denmark/DCGC-16657/2020|EPI_ISL_714897|202 ···················· ···················· ····················· ···················· ···················· ····················
X 457 hCoV-19/Denmark/DCGC-16658/2020|EPI_ISL_714898|202 ···················· ···················· ····················· ···················· ···················· ····················
X 458 hCoV-19/Denmark/DCGC-16661/2020|EPI_ISL_714899|202 ···················· ···················· ····················· ···················· ···················· ····················
X 459 hCoV-19/England/CAMC-C45ECD/2020|EPI_ISL_718390|20 ···················· ···················· ····················· ···················· ······T············· ····················
X 460 hCoV-19/England/CAMC-C47AB8/2020|EPI_ISL_718393|20 ···················· ···················· ····················· ···················· ···················· ····················
X 461 hCoV-19/England/CAMC-C45EDC/2020|EPI_ISL_718397|20 ···················· ···················· ····················· ···················· ······T············· ····················
X 462 hCoV-19/England/CAMC-BBCD89/2020|EPI_ISL_724201|20 ···················· ···················· ····················· ···················· ···················· ····················
X 463 hCoV-19/England/CAMC-BBCD98/2020|EPI_ISL_724202|20 ···················· ···················· ····················· ···················· ···················· ····················
X 464 hCoV-19/England/CAMC-BBCDA7/2020|EPI_ISL_724203|20 ···················· ···················· ····················· ···················· ···················· ····················
X 465 hCoV-19/England/CAMC-BBCDB6/2020|EPI_ISL_724204|20 ···················· ···················· ····················· ···················· ···················· ····················
X 466 hCoV-19/England/CAMC-BBCDC5/2020|EPI_ISL_724205|20 ···················· ···················· ····················· ···················· ···················· ····················
X 467 hCoV-19/England/CAMC-BBCE86/2020|EPI_ISL_724206|20 ···················· ···················· ····················· ···················· ···················· ····················
X 468 hCoV-19/England/CAMC-BBCEB3/2020|EPI_ISL_724207|20 ···················· ···················· ····················· ···················· ···················· ····················
X 469 hCoV-19/England/CAMC-BBCEC2/2020|EPI_ISL_724208|20 ···················· ···················· ····················· ···················· ···················· ····················
X 470 hCoV-19/England/CAMC-BBCED1/2020|EPI_ISL_724209|20 ···················· ···················· ····················· ···················· ···················· ····················
X 471 hCoV-19/Scotland/CVR6062/2020|EPI_ISL_724211|2020- ···················· ···················· ····················· ···················· ···················· ····················
X 472 hCoV-19/Scotland/CVR6073/2020|EPI_ISL_724212|2020- ···················· ···················· ····················· ···················· ···················· ····················
X 473 hCoV-19/Scotland/CVR6080/2020|EPI_ISL_724213|2020- ···················· ···················· ····················· ···················· ···················· ····················
X 474 hCoV-19/Scotland/CVR6120/2020|EPI_ISL_724214|2020- ···················· ···················· ····················· ···················· ···················· ····················
X 475 hCoV-19/Scotland/CVR6121/2020|EPI_ISL_724215|2020- ···················· ···················· ····················· ···················· ···················· ····················
X 476 hCoV-19/Scotland/CVR6122/2020|EPI_ISL_724216|2020- ···················· ···················· ····················· ···················· ···················· ····················
X 477 hCoV-19/Scotland/CVR6128/2020|EPI_ISL_724219|2020- ···················· ···················· ····················· ···················· ···················· ····················
X 478 hCoV-19/Scotland/CVR6129/2020|EPI_ISL_724220|2020- ···················· ···················· ····················· ···················· ···················· ····················
X 479 hCoV-19/Scotland/CVR6134/2020|EPI_ISL_724221|2020- ···················· ·A·················· ····················· ···················· ···················· ··A·················
X 480 hCoV-19/Scotland/EDB10322/2020|EPI_ISL_724223|2020 ···················· ···················· ····················· ···················· ···················· ····················
X 481 hCoV-19/Scotland/EDB10367/2020|EPI_ISL_724224|2020 ···················· ···················· ····················· ···················· ···················· ····················
X 482 hCoV-19/Scotland/EDB10410/2020|EPI_ISL_724225|2020 ···················· ···················· ····················· ···················· ···················· ····················
X 483 hCoV-19/Scotland/EDB10522/2020|EPI_ISL_724226|2020 ···················· ···················· ····················· ···················· ···················· ····················

| | | | | | | | |
|---|---|---|---|---|---|---|---|
| X | 484 hCoV-19/Scotland/EDB10745/2020\|EPI_ISL_724227\|2020 | ···················· | ···················· | ···················· | ···················· | ···················· | ···················· |
| X | 485 hCoV-19/Scotland/EDB10767/2020\|EPI_ISL_724228\|2020 | ···················· | ···················· | ···················· | ···················· | ···················· | ···················· |
| X | 486 hCoV-19/Scotland/EDB10768/2020\|EPI_ISL_724229\|2020 | ···················· | ···················· | ···················· | ···················· | ···················· | ···················· |
| X | 487 hCoV-19/Scotland/EDB10771/2020\|EPI_ISL_724230\|2020 | ···················· | ···················· | ···················· | ···················· | ···················· | ···················· |
| X | 488 hCoV-19/Scotland/EDB10773/2020\|EPI_ISL_724231\|2020 | ···················· | ···················· | ···················· | ···················· | ···················· | ···················· |
| X | 489 hCoV-19/Scotland/EDB10775/2020\|EPI_ISL_724232\|2020 | ···················· | ···················· | ···················· | ···················· | ···················· | ···················· |
| X | 490 hCoV-19/Scotland/EDB10777/2020\|EPI_ISL_724233\|2020 | ···················· | ···················· | ···················· | ···················· | ···················· | ···················· |
| X | 491 hCoV-19/Scotland/EDB10781/2020\|EPI_ISL_724235\|2020 | ···················· | ···················· | ···················· | ···················· | ···················· | ···················· |
| X | 492 hCoV-19/Scotland/EDB10785/2020\|EPI_ISL_724237\|2020 | ···················· | ···················· | ···················· | ···················· | ···················· | ···················· |
| X | 493 hCoV-19/Scotland/EDB10799/2020\|EPI_ISL_724238\|2020 | ···················· | ···················· | ···················· | ···················· | ···················· | ···················· |
| X | 494 hCoV-19/Scotland/EDB10803/2020\|EPI_ISL_724239\|2020 | ···················· | ···················· | ···················· | ···················· | ···················· | ···················· |
| X | 496 hCoV-19/Scotland/EDB10824/2020\|EPI_ISL_724241\|2020 | ···················· | ···················· | ···················· | ···················· | ···················· | ···················· |
| X | 497 hCoV-19/Scotland/EDB10836/2020\|EPI_ISL_724242\|2020 | ···················· | ···················· | ···················· | ···················· | ···················· | ···················· |
| X | 498 hCoV-19/Scotland/EDB10847/2020\|EPI_ISL_724243\|2020 | ···················· | ···················· | ···················· | ···················· | ···················· | ···················· |
| X | 499 hCoV-19/Scotland/EDB10851/2020\|EPI_ISL_724244\|2020 | ···················· | ···················· | ···················· | ···················· | ···················· | ···················· |
| X | 500 hCoV-19/Scotland/EDB10853/2020\|EPI_ISL_724245\|2020 | ···················· | ···················· | ···················· | ···················· | ···················· | ···················· |
| X | 501 hCoV-19/Scotland/EDB10858/2020\|EPI_ISL_724246\|2020 | ···················· | ···················· | ···················· | ···················· | ···················· | ···················· |
| X | 502 hCoV-19/Scotland/EDB10887/2020\|EPI_ISL_724247\|2020 | ···················· | ···················· | ···················· | ···················· | ···················· | ···················· |
| X | 503 hCoV-19/Scotland/EDB10897/2020\|EPI_ISL_724248\|2020 | ···················· | ···················· | ···················· | ···················· | ···················· | ···················· |
| X | 504 hCoV-19/Scotland/EDB10900/2020\|EPI_ISL_724249\|2020 | ···················· | ···················· | ···················· | ···················· | ···················· | ···················· |
| X | 505 hCoV-19/Scotland/EDB10941/2020\|EPI_ISL_724250\|2020 | ···················· | ···················· | ···················· | ···················· | ···················· | ···················· |
| X | 506 hCoV-19/Scotland/EDB10998/2020\|EPI_ISL_724251\|2020 | ···················· | ···················· | ···················· | ···················· | ···················· | ···················· |
| X | 507 hCoV-19/Scotland/EDB11004/2020\|EPI_ISL_724252\|2020 | ···················· | ···················· | ···················· | ···················· | ···················· | ···················· |
| X | 508 hCoV-19/England/EXET-1393F5/2020\|EPI_ISL_724253\|20 | ···················· | ···················· | ···················· | ···················· | ···················· | ···················· |
| X | 509 hCoV-19/England/EXET-13998A/2020\|EPI_ISL_724255\|20 | ···················· | ···················· | ···················· | ···················· | ······T············· | ···················· |
| X | 510 hCoV-19/England/EXET-1399D5/2020\|EPI_ISL_724258\|20 | ···················· | ···················· | ···················· | ···················· | ···················· | ···················· |
| X | 511 hCoV-19/England/EXET-139FF1/2020\|EPI_ISL_724259\|20 | ···················· | ···················· | ···················· | ···················· | ···················· | ···················· |
| X | 512 hCoV-19/England/EXET-13ABFC/2020\|EPI_ISL_724260\|20 | ···················· | ···················· | ···················· | ···················· | ···················· | ···················· |
| X | 513 hCoV-19/Denmark/DCGC-13175/2020\|EPI_ISL_712260\|202 | ···················· | ···················· | ···················· | ···················· | ···················· | ···················· |
| X | 514 hCoV-19/Denmark/DCGC-13252/2020\|EPI_ISL_712261\|202 | ···················· | ···················· | ···················· | ···················· | ···················· | ···················· |
| X | 515 hCoV-19/Denmark/DCGC-13311/2020\|EPI_ISL_712262\|202 | ···················· | ···················· | ···················· | ···················· | ···················· | ···················· |
| X | 516 hCoV-19/Denmark/DCGC-13314/2020\|EPI_ISL_712263\|202 | ···················· | ···················· | ···················· | ···················· | ···················· | ···················· |
| X | 517 hCoV-19/Denmark/DCGC-13322/2020\|EPI_ISL_712264\|202 | ···················· | ···················· | ···················· | ···················· | ···················· | ···················· |
| X | 518 hCoV-19/Denmark/DCGC-13334/2020\|EPI_ISL_712265\|202 | ···················· | ···················· | ···················· | ···················· | ···················· | ···················· |
| X | 519 hCoV-19/Denmark/DCGC-13351/2020\|EPI_ISL_712267\|202 | ···················· | ···················· | ···················· | ···················· | ···················· | ···················· |
| X | 520 hCoV-19/Denmark/DCGC-13838/2020\|EPI_ISL_712290\|202 | ···················· | ···················· | ···················· | ···················· | ···················· | ···················· |
| X | 521 hCoV-19/Denmark/DCGC-13861/2020\|EPI_ISL_712291\|202 | ···················· | ···················· | ···················· | ···················· | ···················· | ···················· |
| X | 522 hCoV-19/Denmark/DCGC-13888/2020\|EPI_ISL_712292\|202 | ···················· | ···················· | ···················· | ···················· | ···················· | ···················· |
| X | 523 hCoV-19/Denmark/DCGC-13896/2020\|EPI_ISL_712293\|202 | ···················· | ···················· | ···················· | ···················· | ···················· | ···················· |
| X | 524 hCoV-19/Denmark/DCGC-13050/2020\|EPI_ISL_712294\|202 | ···················· | ···················· | ···················· | ···················· | ···················· | ···················· |
| X | 525 hCoV-19/Denmark/DCGC-12963/2020\|EPI_ISL_712295\|202 | ···················· | ···················· | ···················· | ···················· | ···················· | ···················· |
| X | 526 hCoV-19/Denmark/DCGC-13402/2020\|EPI_ISL_712296\|202 | ···················· | ···················· | ···················· | ···················· | ···················· | ···················· |
| X | 527 hCoV-19/Denmark/DCGC-13475/2020\|EPI_ISL_712297\|202 | ···················· | ···················· | ···················· | ···················· | ···················· | ···················· |
| X | 528 hCoV-19/Denmark/DCGC-13543/2020\|EPI_ISL_712298\|202 | ···················· | ···················· | ···················· | ···················· | ···················· | ···················· |
| X | 529 hCoV-19/Denmark/DCGC-13731/2020\|EPI_ISL_712299\|202 | ···················· | ···················· | ···················· | ···················· | ···················· | ···················· |
| X | 530 hCoV-19/Denmark/DCGC-15028/2020\|EPI_ISL_713590\|202 | ···················· | ···················· | ···················· | ···················· | ···················· | ···················· |
| X | 531 hCoV-19/Denmark/DCGC-15029/2020\|EPI_ISL_713591\|202 | ···················· | ···················· | ···················· | ···················· | ···················· | ···················· |
| X | 532 hCoV-19/Denmark/DCGC-15030/2020\|EPI_ISL_713592\|202 | ···················· | ···················· | ···················· | ···················· | ···················· | ···················· |
| X | 533 hCoV-19/Denmark/DCGC-15032/2020\|EPI_ISL_713593\|202 | ···················· | ···················· | ···················· | ···················· | ···················· | ···················· |
| X | 534 hCoV-19/Denmark/DCGC-15033/2020\|EPI_ISL_713594\|202 | ···················· | ···················· | ···················· | ···················· | ···················· | ···················· |
| X | 535 hCoV-19/Denmark/DCGC-15035/2020\|EPI_ISL_713595\|202 | ···················· | ···················· | ···················· | ···················· | ···················· | ···················· |
| X | 536 hCoV-19/Denmark/DCGC-15036/2020\|EPI_ISL_713596\|202 | ···················· | ···················· | ···················· | ···················· | ···················· | ···················· |
| X | 537 hCoV-19/Denmark/DCGC-15041/2020\|EPI_ISL_713600\|202 | ···················· | ···················· | ···················· | ···················· | ···················· | ···················· |
| X | 538 hCoV-19/Denmark/DCGC-15044/2020\|EPI_ISL_713601\|202 | ···················· | ···················· | ···················· | ···················· | ···················· | ···················· |
| X | 539 hCoV-19/Denmark/DCGC-15045/2020\|EPI_ISL_713602\|202 | ···················· | ···················· | ···················· | ···················· | ···················· | ···················· |
| X | 540 hCoV-19/Denmark/DCGC-15046/2020\|EPI_ISL_713603\|202 | ···················· | ···················· | ···················· | ···················· | ···················· | ···················· |
| X | 541 hCoV-19/Denmark/DCGC-15047/2020\|EPI_ISL_713604\|202 | ···················· | ···················· | ···················· | ···················· | ···················· | ···················· |
| X | 542 hCoV-19/Denmark/DCGC-15051/2020\|EPI_ISL_713605\|202 | ···················· | ····A··············· | ···················· | ···················· | ···················· | ···················· |
| X | 543 hCoV-19/Denmark/DCGC-15052/2020\|EPI_ISL_713606\|202 | ···················· | ···················· | ···················· | ···················· | ···················· | ···················· |
| X | 544 hCoV-19/Denmark/DCGC-15056/2020\|EPI_ISL_713607\|202 | ···················· | ···················· | ···················· | ···················· | ···················· | ···················· |
| X | 545 hCoV-19/Denmark/DCGC-15057/2020\|EPI_ISL_713608\|202 | ···················· | ···················· | ···················· | ···················· | ···················· | ···················· |
| X | 546 hCoV-19/Denmark/DCGC-15058/2020\|EPI_ISL_713609\|202 | ···················· | ···················· | ···················· | ···················· | ···················· | ···················· |
| X | 547 hCoV-19/Denmark/DCGC-15059/2020\|EPI_ISL_713610\|202 | ···················· | ···················· | ···················· | ···················· | ···················· | ···················· |
| X | 548 hCoV-19/Denmark/DCGC-15063/2020\|EPI_ISL_713611\|202 | ···················· | ···················· | ···················· | ···················· | ···················· | ···················· |
| X | 549 hCoV-19/Denmark/DCGC-15064/2020\|EPI_ISL_713612\|202 | ···················· | ···················· | ···················· | ···················· | ···················· | ···················· |
| X | 550 hCoV-19/Denmark/DCGC-15065/2020\|EPI_ISL_713613\|202 | ···················· | ···················· | ···················· | ···················· | ···················· | ···················· |
| X | 551 hCoV-19/Denmark/DCGC-15067/2020\|EPI_ISL_713614\|202 | ···················· | ···················· | ···················· | ···················· | ···················· | ···················· |
| X | 552 hCoV-19/Denmark/DCGC-15068/2020\|EPI_ISL_713615\|202 | ···················· | ···················· | ···················· | ···················· | ···················· | ···················· |
| X | 553 hCoV-19/Denmark/DCGC-15113/2020\|EPI_ISL_713638\|202 | ···················· | ···················· | ···················· | ···················· | ···················· | ···················· |
| X | 554 hCoV-19/Denmark/DCGC-15114/2020\|EPI_ISL_713639\|202 | ···················· | ····A··············· | ···················· | ···················· | ···················· | ···················· |
| X | 555 hCoV-19/Denmark/DCGC-15129/2020\|EPI_ISL_713645\|202 | ···················· | ···················· | ···················· | ···················· | ···················· | ···················· |
| X | 556 hCoV-19/Denmark/DCGC-15130/2020\|EPI_ISL_713646\|202 | ···················· | ···················· | ···················· | ···················· | ···················· | ···················· |
| X | 557 hCoV-19/Denmark/DCGC-15131/2020\|EPI_ISL_713647\|202 | ···················· | ·A·················· | ···················· | ···················· | ···················· | ···A················ |
| X | 558 hCoV-19/Denmark/DCGC-15132/2020\|EPI_ISL_713648\|202 | ···················· | ···················· | ···················· | ···················· | ······T············· | ···················· |
| X | 559 hCoV-19/Denmark/DCGC-16662/2020\|EPI_ISL_714900\|202 | ···················· | ···················· | ···················· | ···················· | ···················· | ···················· |
| X | 560 hCoV-19/Denmark/DCGC-16663/2020\|EPI_ISL_714901\|202 | ···················· | ···················· | ···················· | ···················· | ···················· | ···················· |
| X | 561 hCoV-19/Denmark/DCGC-16664/2020\|EPI_ISL_714902\|202 | ···················· | ···················· | ···················· | ···················· | ···················· | ···················· |
| X | 562 hCoV-19/Denmark/DCGC-16667/2020\|EPI_ISL_714903\|202 | ···················· | ···················· | ···················· | ···················· | ···················· | ···················· |
| X | 563 hCoV-19/Denmark/DCGC-16670/2020\|EPI_ISL_714904\|202 | ···················· | ···················· | ···················· | ···················· | ···················· | ···················· |
| X | 564 hCoV-19/Denmark/DCGC-16672/2020\|EPI_ISL_714905\|202 | ···················· | ···················· | ···················· | ···················· | ···················· | ···················· |

| | | | | | | | |
|---|---|---|---|---|---|---|---|
| X | 565 hCoV-19/Denmark/DCGC-16674/2020\|EPI_ISL_714906\|202 | ···················· | ···················· | ···················· | ···················· | ···················· | ···················· |
| X | 566 hCoV-19/Denmark/DCGC-16676/2020\|EPI_ISL_714907\|202 | ···················· | ····A··············· | ···················· | ···················· | ···················· | ···················· |
| X | 567 hCoV-19/Denmark/DCGC-16677/2020\|EPI_ISL_714908\|202 | ···················· | ·A·················· | ···················· | ···················· | ···················· | ···················· |
| X | 568 hCoV-19/Denmark/DCGC-16678/2020\|EPI_ISL_714909\|202 | ···················· | ···················· | ···················· | ···················· | ···················· | ···················· |
| X | 569 hCoV-19/Denmark/DCGC-16680/2020\|EPI_ISL_714910\|202 | ···················· | ···················· | ···················· | ···················· | ···················· | ···················· |
| X | 570 hCoV-19/Denmark/DCGC-16681/2020\|EPI_ISL_714911\|202 | ···················· | ···················· | ···················· | ···················· | ···················· | ···················· |
| X | 571 hCoV-19/Denmark/DCGC-16682/2020\|EPI_ISL_714912\|202 | ···················· | ···················· | ···················· | ···················· | ···················· | ···················· |
| X | 572 hCoV-19/Denmark/DCGC-16684/2020\|EPI_ISL_714913\|202 | ···················· | ···················· | ···················· | ···················· | ···················· | ···················· |
| X | 573 hCoV-19/Denmark/DCGC-16685/2020\|EPI_ISL_714914\|202 | ···················· | ···················· | ···················· | ···················· | ···················· | ···················· |
| X | 574 hCoV-19/Denmark/DCGC-16686/2020\|EPI_ISL_714915\|202 | ···················· | ···················· | ···················· | ···················· | ···················· | ···················· |
| X | 575 hCoV-19/Denmark/DCGC-16687/2020\|EPI_ISL_714916\|202 | ···················· | ···················· | ···················· | ···················· | ···················· | ···················· |
| X | 576 hCoV-19/Denmark/DCGC-16688/2020\|EPI_ISL_714917\|202 | ···················· | ···················· | ···················· | ···················· | ···················· | ···················· |
| X | 577 hCoV-19/Denmark/DCGC-16689/2020\|EPI_ISL_714918\|202 | ···················· | ···················· | ···················· | ···················· | ···················· | ···················· |
| X | 578 hCoV-19/Denmark/DCGC-16690/2020\|EPI_ISL_714919\|202 | ···················· | ···················· | ···················· | ···················· | ···················· | ···················· |
| X | 579 hCoV-19/Denmark/DCGC-16691/2020\|EPI_ISL_714920\|202 | ···················· | ···················· | ···················· | ···················· | ···················· | ···················· |
| X | 580 hCoV-19/Denmark/DCGC-16692/2020\|EPI_ISL_714921\|202 | ···················· | ···················· | ···················· | ···················· | ···················· | ···················· |
| X | 581 hCoV-19/Denmark/DCGC-16693/2020\|EPI_ISL_714922\|202 | ···················· | ·A·················· | ···················· | ···················· | ···················· | ···················· |
| X | 582 hCoV-19/Denmark/DCGC-16696/2020\|EPI_ISL_714923\|202 | ···················· | ···················· | ···················· | ···················· | ···················· | ···················· |
| X | 583 hCoV-19/Denmark/DCGC-16702/2020\|EPI_ISL_714924\|202 | ···················· | ···················· | ···················· | ···················· | ···················· | ···················· |
| X | 584 hCoV-19/Denmark/DCGC-16703/2020\|EPI_ISL_714925\|202 | ···················· | ···················· | ···················· | ···················· | ···················· | ···················· |
| X | 585 hCoV-19/Denmark/DCGC-16704/2020\|EPI_ISL_714926\|202 | ···················· | ····A··············· | ···················· | ···················· | ···················· | ···················· |
| X | 586 hCoV-19/Denmark/DCGC-16705/2020\|EPI_ISL_714927\|202 | ···················· | ···················· | ···················· | ···················· | ···················· | ···················· |
| X | 587 hCoV-19/Denmark/DCGC-16710/2020\|EPI_ISL_714928\|202 | ···················· | ···················· | ···················· | ···················· | ···················· | ···················· |
| X | 588 hCoV-19/Denmark/DCGC-16732/2020\|EPI_ISL_714929\|202 | ···················· | ···················· | ···················· | ···················· | ···················· | ···················· |
| X | 589 hCoV-19/Denmark/DCGC-16733/2020\|EPI_ISL_714930\|202 | ···················· | ···················· | ···················· | ···················· | ···················· | ···················· |
| X | 590 hCoV-19/Denmark/DCGC-16734/2020\|EPI_ISL_714931\|202 | ···················· | ···················· | ···················· | ···················· | ···················· | ···················· |
| X | 591 hCoV-19/Denmark/DCGC-16737/2020\|EPI_ISL_714932\|202 | ···················· | ···················· | ···················· | ···················· | ···················· | ···················· |
| X | 592 hCoV-19/Denmark/DCGC-16739/2020\|EPI_ISL_714933\|202 | ···················· | ···················· | ···················· | ···················· | ···················· | ···················· |
| X | 593 hCoV-19/Denmark/DCGC-16740/2020\|EPI_ISL_714934\|202 | ···················· | ···················· | ···················· | ···················· | ···················· | ···················· |
| X | 594 hCoV-19/Denmark/DCGC-16741/2020\|EPI_ISL_714935\|202 | ···················· | ···················· | ···················· | ···················· | ···················· | ···················· |
| X | 595 hCoV-19/Denmark/DCGC-16742/2020\|EPI_ISL_714936\|202 | ···················· | ···················· | ···················· | ···················· | ···················· | ···················· |
| X | 596 hCoV-19/Denmark/DCGC-16748/2020\|EPI_ISL_714937\|202 | ···················· | ···················· | ···················· | ···················· | ···················· | ···················· |
| X | 597 hCoV-19/England/CAMC-C453BF/2020\|EPI_ISL_718411\|20 | ···················· | ···················· | ···················· | ···················· | ···················· | ···················· |
| X | 598 hCoV-19/England/CAMC-C44286/2020\|EPI_ISL_718415\|20 | ···················· | ···················· | ···················· | ···················· | ···················· | ···················· |
| X | 599 hCoV-19/England/CAMC-C46952/2020\|EPI_ISL_718416\|20 | ···················· | ···················· | ···················· | ···················· | ···················· | ···················· |
| X | 600 hCoV-19/England/CAMC-C40536/2020\|EPI_ISL_718417\|20 | ···················· | ···················· | ···················· | ···················· | ···················· | ···················· |
| X | 601 hCoV-19/England/CAMC-C459EA/2020\|EPI_ISL_718423\|20 | ···················· | ···················· | ···················· | ···················· | ···················· | ···················· |
| X | 602 hCoV-19/England/CAMC-C45F9D/2020\|EPI_ISL_718424\|20 | ···················· | ···················· | ···················· | ···················· | ···················· | ···················· |
| X | 603 hCoV-19/England/CAMC-C44D0D/2020\|EPI_ISL_718434\|20 | ···················· | ···················· | ···················· | ···················· | ···················· | ···················· |
| X | 604 hCoV-19/England/CAMC-C45B8A/2020\|EPI_ISL_718436\|20 | ···················· | ···················· | ···················· | ···················· | ···················· | ···················· |
| X | 605 hCoV-19/England/CAMC-C3FCDB/2020\|EPI_ISL_718441\|20 | ···················· | ·A·················· | ···················· | ···················· | ···················· | ···················· |
| X | 606 hCoV-19/England/CAMC-C46749/2020\|EPI_ISL_718448\|20 | ···················· | ···················· | ···················· | ···················· | ···················· | ···················· |
| X | 607 hCoV-19/England/CAMC-C44DD0/2020\|EPI_ISL_718449\|20 | ···················· | ···················· | ···················· | ···················· | ···················· | ···················· |
| X | 608 hCoV-19/England/CAMC-C46B5C/2020\|EPI_ISL_718450\|20 | ···················· | ···················· | ···················· | ···················· | ···················· | ···················· |
| X | 609 hCoV-19/England/CAMC-C4699E/2020\|EPI_ISL_718451\|20 | ···················· | ···················· | ···················· | ···················· | ···················· | ···················· |
| X | 610 hCoV-19/England/CAMC-C45B99/2020\|EPI_ISL_718453\|20 | ···················· | ···················· | ···················· | ···················· | ···················· | ···················· |
| X | 611 hCoV-19/England/CAMC-C46A6E/2020\|EPI_ISL_718454\|20 | ···················· | ···················· | ···················· | ···················· | ···················· | ···················· |
| X | 612 hCoV-19/England/CAMC-C44D49/2020\|EPI_ISL_718467\|20 | ···················· | ···················· | ···················· | ···················· | ···················· | ···················· |
| X | 613 hCoV-19/England/CAMC-C46D29/2020\|EPI_ISL_718468\|20 | ···················· | ···················· | ···················· | ···················· | ···················· | ···················· |
| X | 614 hCoV-19/England/CAMC-C4672B/2020\|EPI_ISL_718472\|20 | ···················· | ···················· | ···················· | ···················· | ······T············· | ···················· |
| X | 615 hCoV-19/England/CAMC-C40369/2020\|EPI_ISL_718473\|20 | ···················· | ···················· | ···················· | ···················· | ···················· | ···················· |
| X | 616 hCoV-19/England/CAMC-C44DFE/2020\|EPI_ISL_718475\|20 | ···················· | ···················· | ···················· | ···················· | ···················· | ···················· |
| X | 617 hCoV-19/England/CAMC-C3F9F2/2020\|EPI_ISL_718476\|20 | ···················· | ···················· | ···················· | ···················· | ···················· | ···················· |
| X | 618 hCoV-19/England/CAMC-C4698F/2020\|EPI_ISL_718477\|20 | ···················· | ···················· | ···················· | ···················· | ···················· | ···················· |
| X | 619 hCoV-19/England/CAMC-C45AAB/2020\|EPI_ISL_718478\|20 | ···················· | ···················· | ···················· | ···················· | ···················· | ···················· |
| X | 620 hCoV-19/England/CAMC-C46B3E/2020\|EPI_ISL_718482\|20 | ···················· | ···················· | ···················· | ···················· | ···················· | ···················· |
| X | 621 hCoV-19/England/CAMC-C44DA3/2020\|EPI_ISL_718484\|20 | ···················· | ···················· | ···················· | ···················· | ···················· | ···················· |
| X | 622 hCoV-19/England/CAMC-C3F998/2020\|EPI_ISL_718492\|20 | ···················· | ···················· | ···················· | ···················· | ···················· | ···················· |
| X | 623 hCoV-19/England/CAMC-C479E8/2020\|EPI_ISL_718493\|20 | ···················· | ···················· | ···················· | ···················· | ···················· | ···················· |
| X | 624 hCoV-19/England/CAMC-C45F24/2020\|EPI_ISL_718495\|20 | ···················· | ···················· | ···················· | ···················· | ···················· | ···················· |
| X | 625 hCoV-19/England/CAMC-C46679/2020\|EPI_ISL_718496\|20 | ···················· | ···················· | ···················· | ···················· | ···················· | ···················· |
| X | 626 hCoV-19/England/CAMC-C4607B/2020\|EPI_ISL_718497\|20 | ···················· | ···················· | ···················· | ···················· | ···················· | ···················· |
| X | 627 hCoV-19/England/QEUH-BA1051/2020\|EPI_ISL_719700\|20 | ···················· | ·A·················· | ···················· | ···················· | ···················· | ···················· |
| X | 628 hCoV-19/England/QEUH-BA1A53/2020\|EPI_ISL_719701\|20 | ···················· | ···················· | ···················· | ···················· | ···················· | ···················· |
| X | 629 hCoV-19/England/QEUH-BA1938/2020\|EPI_ISL_719702\|20 | ···················· | ···················· | ···················· | ···················· | ···················· | ···················· |
| X | 630 hCoV-19/England/QEUH-BA1992/2020\|EPI_ISL_719703\|20 | ···················· | ···················· | ···················· | ···················· | ···················· | ···················· |
| X | 631 hCoV-19/England/QEUH-BA1974/2020\|EPI_ISL_719705\|20 | ···················· | ···················· | ···················· | ···················· | ···················· | ···················· |
| X | 632 hCoV-19/England/QEUH-BA0AEB/2020\|EPI_ISL_719707\|20 | ···················· | ···················· | ···················· | ···················· | ···················· | ···················· |
| X | 633 hCoV-19/England/QEUH-B9E4F1/2020\|EPI_ISL_719708\|20 | ···················· | ···················· | ···················· | ···················· | ······T············· | ···················· |
| X | 634 hCoV-19/England/QEUH-BA075D/2020\|EPI_ISL_719709\|20 | ···················· | ···················· | ···················· | ···················· | ···················· | ···················· |
| X | 635 hCoV-19/England/QEUH-B9EF87/2020\|EPI_ISL_719710\|20 | ···················· | ···················· | ···················· | ···················· | ···················· | ···················· |
| X | 636 hCoV-19/England/QEUH-BA0ADC/2020\|EPI_ISL_719712\|20 | ···················· | ···················· | ···················· | ···················· | ···················· | ···················· |
| X | 637 hCoV-19/England/QEUH-BA04DE/2020\|EPI_ISL_719713\|20 | ···················· | ···················· | ···················· | ···················· | ···················· | ···················· |
| X | 638 hCoV-19/England/QEUH-BA03E0/2020\|EPI_ISL_719714\|20 | ···················· | ···················· | ···················· | ···················· | ···················· | ···················· |
| X | 639 hCoV-19/England/QEUH-BA191A/2020\|EPI_ISL_719715\|20 | ···················· | ···················· | ···················· | ···················· | ···················· | ···················· |
| X | 640 hCoV-19/England/QEUH-BA00AD/2020\|EPI_ISL_719716\|20 | ···················· | ···················· | ···················· | ···················· | ···················· | ···················· |
| X | 641 hCoV-19/England/QEUH-B9EF69/2020\|EPI_ISL_719717\|20 | ···················· | ···················· | ···················· | ···················· | ···················· | ···················· |
| X | 642 hCoV-19/England/QEUH-BA03A4/2020\|EPI_ISL_719720\|20 | ···················· | ···················· | ···················· | ···················· | ···················· | ···················· |
| X | 643 hCoV-19/England/QEUH-BA0614/2020\|EPI_ISL_719722\|20 | ···················· | ·A·················· | ···················· | ···················· | ···················· | ···A················ |
| X | 644 hCoV-19/England/QEUH-BA0799/2020\|EPI_ISL_719723\|20 | ···················· | ···················· | ···················· | ···················· | ···················· | ···················· |

| | | | | | | | |
|---|---|---|---|---|---|---|---|
| X | 645 hCoV-19/England/QEUH-BA0A72/2020\|EPI_ISL_719724\|20 | ···················· | ···················· | ···················· | ···················· | ···················· | ···················· |
| X | 646 hCoV-19/England/QEUH-BA040B/2020\|EPI_ISL_719725\|20 | ···················· | ···················· | ···················· | ···················· | ···················· | ···················· |
| X | 647 hCoV-19/England/QEUH-BA08C3/2020\|EPI_ISL_719726\|20 | ···················· | ···················· | ···················· | ···················· | ···················· | ···················· |
| X | 649 hCoV-19/England/QEUH-B9FF68/2020\|EPI_ISL_719728\|20 | ···················· | ···················· | ···················· | ···················· | ···················· | ···················· |
| X | 650 hCoV-19/England/QEUH-B9FF59/2020\|EPI_ISL_719729\|20 | ···················· | ···················· | ···················· | ···················· | ···················· | ···················· |
| X | 651 hCoV-19/England/QEUH-BA0429/2020\|EPI_ISL_719730\|20 | ···················· | ···················· | ···················· | ···················· | ···················· | ···················· |
| X | 652 hCoV-19/England/QEUH-BA0113/2020\|EPI_ISL_719731\|20 | ···················· | ···················· | ···················· | ···················· | ···················· | ···················· |
| X | 653 hCoV-19/England/QEUH-BA0517/2020\|EPI_ISL_719732\|20 | ···················· | ···················· | ···················· | ···················· | ···················· | ···················· |
| X | 654 hCoV-19/England/QEUH-BA019B/2020\|EPI_ISL_719733\|20 | ···················· | ···················· | ···················· | ···················· | ···················· | ···················· |
| X | 656 hCoV-19/England/QEUH-BA172F/2020\|EPI_ISL_719735\|20 | ···················· | ···················· | ···················· | ···················· | ···················· | ···················· |
| X | 657 hCoV-19/England/QEUH-BA1983/2020\|EPI_ISL_719736\|20 | ···················· | ···················· | ···················· | ···················· | ···················· | ···················· |
| X | 658 hCoV-19/England/QEUH-B9EF78/2020\|EPI_ISL_719737\|20 | ···················· | ···················· | ···················· | ···················· | ···················· | ···················· |
| X | 659 hCoV-19/England/QEUH-BA03D1/2020\|EPI_ISL_719738\|20 | ···················· | ···················· | ···················· | ···················· | ···················· | ···················· |
| X | 660 hCoV-19/England/QEUH-BA1042/2020\|EPI_ISL_719739\|20 | ···················· | ···················· | ···················· | ···················· | ···················· | ···················· |
| X | 661 hCoV-19/England/QEUH-BA0456/2020\|EPI_ISL_719740\|20 | ···················· | ···················· | ···················· | ···················· | ···················· | ···················· |
| X | 662 hCoV-19/England/QEUH-BA0007/2020\|EPI_ISL_719742\|20 | ···················· | ···················· | ···················· | ···················· | ···················· | ···················· |
| X | 663 hCoV-19/England/QEUH-BA0034/2020\|EPI_ISL_719744\|20 | ···················· | ···················· | ···················· | ···················· | ···················· | ···················· |
| X | 664 hCoV-19/England/QEUH-BA16D7/2020\|EPI_ISL_719748\|20 | ···················· | ···················· | ···················· | ···················· | ···················· | ···················· |
| X | 665 hCoV-19/England/QEUH-BA00E9/2020\|EPI_ISL_719749\|20 | ···················· | ···················· | ···················· | ···················· | ···················· | ···················· |
| X | 666 hCoV-19/England/QEUH-BA04FC/2020\|EPI_ISL_719750\|20 | ···················· | ···················· | ···················· | ···················· | ···················· | ···················· |
| X | 667 hCoV-19/England/QEUH-BA19B0/2020\|EPI_ISL_719752\|20 | ···················· | ·A·················· | ···················· | ···················· | ···················· | ···················· |
| X | 668 hCoV-19/England/QEUH-BA017D/2020\|EPI_ISL_719753\|20 | ···················· | ·A·················· | ···················· | ···················· | ···················· | ··A················· |
| X | 669 hCoV-19/England/QEUH-BA0DF1/2020\|EPI_ISL_719755\|20 | ···················· | ···················· | ···················· | ···················· | ···················· | ···················· |
| X | 670 hCoV-19/England/QEUH-B9EC35/2020\|EPI_ISL_719756\|20 | ···················· | ···················· | ···················· | ···················· | ···················· | ···················· |
| X | 671 hCoV-19/England/QEUH-BA175C/2020\|EPI_ISL_719758\|20 | ···················· | ···················· | ···················· | ···················· | ···················· | ···················· |
| X | 672 hCoV-19/England/QEUH-BA0016/2020\|EPI_ISL_719759\|20 | ···················· | ···················· | ···················· | ···················· | ···················· | ···················· |
| X | 673 hCoV-19/England/QEUH-BA059F/2020\|EPI_ISL_719760\|20 | ···················· | ···················· | ···················· | ···················· | ···················· | ···················· |
| X | 674 hCoV-19/England/QEUH-B9FF95/2020\|EPI_ISL_719761\|20 | ···················· | ···················· | ···················· | ···················· | ···················· | ···················· |
| X | 675 hCoV-19/England/QEUH-BA0447/2020\|EPI_ISL_719763\|20 | ···················· | ···················· | ···················· | ···················· | ···················· | ···················· |
| X | 676 hCoV-19/England/QEUH-BA0580/2020\|EPI_ISL_719764\|20 | ···················· | ···················· | ···················· | ···················· | ···················· | ···················· |
| X | 677 hCoV-19/England/QEUH-B9EED5/2020\|EPI_ISL_719766\|20 | ···················· | ···················· | ···················· | ···················· | ···················· | ···················· |
| X | 678 hCoV-19/England/QEUH-B9FFA4/2020\|EPI_ISL_719768\|20 | ···················· | ···················· | ···················· | ···················· | ···················· | ···················· |
| X | 679 hCoV-19/England/QEUH-BA0623/2020\|EPI_ISL_719769\|20 | ···················· | ·A·················· | ···················· | ···················· | ···················· | ··A················· |
| X | 680 hCoV-19/England/QEUH-BA078A/2020\|EPI_ISL_719770\|20 | ···················· | ···················· | ···················· | ···················· | ···················· | ···················· |
| X | 681 hCoV-19/England/QEUH-BA1947/2020\|EPI_ISL_719771\|20 | ···················· | ·A·················· | ···················· | ···················· | ···················· | ··A················· |
| X | 682 hCoV-19/England/QEUH-BA19FC/2020\|EPI_ISL_719772\|20 | ···················· | ···················· | ···················· | ···················· | ···················· | ···················· |
| X | 683 hCoV-19/England/QEUH-B9E558/2020\|EPI_ISL_719773\|20 | ···················· | ···················· | ···················· | ···················· | ···················· | ···················· |
| X | 684 hCoV-19/England/QEUH-BA19DE/2020\|EPI_ISL_719774\|20 | ···················· | ···················· | ···················· | ···················· | ···················· | ···················· |
| X | 685 hCoV-19/England/QEUH-B9EF1E/2020\|EPI_ISL_719775\|20 | ···················· | ···················· | ···················· | ···················· | ···················· | ···················· |
| X | 686 hCoV-19/England/QEUH-B9E4C4/2020\|EPI_ISL_719777\|20 | ···················· | ···················· | ···················· | ···················· | ···················· | ···················· |
| X | 687 hCoV-19/England/QEUH-BA06F6/2020\|EPI_ISL_719780\|20 | ···················· | ···················· | ···················· | ···················· | ···················· | ···················· |
| X | 688 hCoV-19/England/QEUH-BA0D00/2020\|EPI_ISL_719781\|20 | ···················· | ···················· | ···················· | ···················· | ···················· | ···················· |
| X | 689 hCoV-19/England/QEUH-BA16F5/2020\|EPI_ISL_719782\|20 | ···················· | ·A·················· | ···················· | ···················· | ···················· | ···················· |
| X | 690 hCoV-19/England/QEUH-B9EBED/2020\|EPI_ISL_719783\|20 | ···················· | ···················· | ···················· | ···················· | ···················· | ···················· |
| X | 691 hCoV-19/England/QEUH-BA1A26/2020\|EPI_ISL_719786\|20 | ···················· | ···················· | ···················· | ···················· | ···················· | ···················· |
| X | 692 hCoV-19/England/QEUH-BA0465/2020\|EPI_ISL_719787\|20 | ···················· | ···················· | ···················· | ···················· | ···················· | ···················· |
| X | 693 hCoV-19/England/QEUH-BA05DB/2020\|EPI_ISL_719788\|20 | ···················· | ···················· | ···················· | ···················· | ···················· | ···················· |
| X | 694 hCoV-19/England/QEUH-BA0720/2020\|EPI_ISL_719789\|20 | ···················· | ···················· | ···················· | ···················· | ···················· | ···················· |
| X | 695 hCoV-19/England/QEUH-B9FFE0/2020\|EPI_ISL_719790\|20 | ···················· | ···················· | ···················· | ···················· | ···················· | ···················· |
| X | 696 hCoV-19/England/QEUH-BA168C/2020\|EPI_ISL_719791\|20 | ···················· | ···················· | ···················· | ···················· | ···················· | ···················· |
| X | 697 hCoV-19/England/QEUH-BA074E/2020\|EPI_ISL_719792\|20 | ···················· | ···················· | ···················· | ···················· | ···················· | ···················· |
| X | 698 hCoV-19/England/QEUH-BA0641/2020\|EPI_ISL_719793\|20 | ···················· | ··············C····· | ···················· | ···················· | ···················· | ···················· |
| X | 699 hCoV-19/England/QEUH-B9E45B/2020\|EPI_ISL_719796\|20 | ···················· | ···················· | ···················· | ···················· | ···················· | ···················· |
| X | 700 hCoV-19/England/QEUH-BA0562/2020\|EPI_ISL_719798\|20 | ···················· | ···················· | ···················· | ···················· | ···················· | ···················· |
| X | 701 hCoV-19/England/MILK-C006B7/2020\|EPI_ISL_720701\|20 | ···················· | ···················· | ···················· | ···················· | ···················· | ···················· |
| X | 702 hCoV-19/England/QEUH-C013EC/2020\|EPI_ISL_720704\|20 | ···················· | ···················· | ···················· | ···················· | ···················· | ···················· |
| X | 703 hCoV-19/England/MILK-BFCC17/2020\|EPI_ISL_720706\|20 | ···················· | ···················· | ···················· | ···················· | ···················· | ···················· |
| X | 704 hCoV-19/England/MILK-BFD584/2020\|EPI_ISL_720707\|20 | ···················· | ···················· | ···················· | ···················· | ······T············· | ···················· |
| X | 705 hCoV-19/England/ALDP-C049C9/2020\|EPI_ISL_720709\|20 | ···················· | ·A·················· | ···················· | ···················· | ···················· | ··A················· |
| X | 706 hCoV-19/England/MILK-BFCD6F/2020\|EPI_ISL_720710\|20 | ···················· | ···················· | ···················· | ···················· | ···················· | ···················· |
| X | 707 hCoV-19/England/ALDP-C049F6/2020\|EPI_ISL_720711\|20 | ···················· | ···················· | ···················· | ···················· | ···················· | ···················· |
| X | 708 hCoV-19/England/MILK-BFF5DD/2020\|EPI_ISL_720712\|20 | ···················· | ···················· | ···················· | ···················· | ···················· | ···················· |
| X | 709 hCoV-19/England/ALDP-C05CCF/2020\|EPI_ISL_720713\|20 | ···················· | ···················· | ···················· | ···················· | ···················· | ···················· |
| X | 710 hCoV-19/England/MILK-BFCBDE/2020\|EPI_ISL_720714\|20 | ···················· | ···················· | ···················· | ···················· | ···················· | ···················· |
| X | 711 hCoV-19/England/MILK-C0064E/2020\|EPI_ISL_720717\|20 | ···················· | ···················· | ···················· | ···················· | ···················· | ···················· |
| X | 712 hCoV-19/England/MILK-BFCF78/2020\|EPI_ISL_720719\|20 | ···················· | ···················· | ···················· | ···················· | ···················· | ···················· |
| X | 713 hCoV-19/England/ALDP-C04AE4/2020\|EPI_ISL_720722\|20 | ···················· | ···················· | ···················· | ···················· | ···················· | ···················· |
| X | 714 hCoV-19/England/MILK-BFF8F2/2020\|EPI_ISL_720723\|20 | ···················· | ···················· | ···················· | ···················· | ···················· | ···················· |
| X | 715 hCoV-19/England/MILK-BFCB47/2020\|EPI_ISL_720724\|20 | ···················· | ···················· | ···················· | ···················· | ···················· | ···················· |
| X | 716 hCoV-19/England/MILK-C008C0/2020\|EPI_ISL_720725\|20 | ···················· | ···················· | ···················· | ···················· | ···················· | ···················· |
| X | 717 hCoV-19/England/QEUH-C03E9E/2020\|EPI_ISL_720726\|20 | ···················· | ···················· | ···················· | ···················· | ···················· | ···················· |
| X | 718 hCoV-19/England/QEUH-C01337/2020\|EPI_ISL_720728\|20 | ···················· | ···················· | ···················· | ···················· | ···················· | ···················· |
| X | 719 hCoV-19/England/MILK-C0063F/2020\|EPI_ISL_720730\|20 | ···················· | ···················· | ···················· | ···················· | ···················· | ···················· |
| X | 720 hCoV-19/England/QEUH-C013DD/2020\|EPI_ISL_720732\|20 | ···················· | ···················· | ···················· | ···················· | ···················· | ···················· |
| X | 721 hCoV-19/England/QEUH-C03EBC/2020\|EPI_ISL_720733\|20 | ···················· | ···················· | ···················· | ···················· | ···················· | ···················· |
| X | 722 hCoV-19/England/MILK-BFF4EF/2020\|EPI_ISL_720736\|20 | ···················· | ···················· | ···················· | ···················· | ···················· | ···················· |
| X | 723 hCoV-19/England/QEUH-C01425/2020\|EPI_ISL_720738\|20 | ···················· | ···················· | ···················· | ···················· | ······T············· | ···················· |
| X | 724 hCoV-19/England/QEUH-C01373/2020\|EPI_ISL_720740\|20 | ···················· | ···················· | ···················· | ···················· | ···················· | ···················· |
| X | 725 hCoV-19/England/ALDP-C04A4E/2020\|EPI_ISL_720741\|20 | ···················· | ···················· | ···················· | ···················· | ···················· | ···················· |
| X | 726 hCoV-19/England/MILK-BFCC26/2020\|EPI_ISL_720742\|20 | ···················· | ···················· | ···················· | ···················· | ···················· | ···················· |

| | | | | | | | | |
|---|---|---|---|---|---|---|---|---|
| X | 727 | hCoV-19/England/ALDP-C049BA/2020\|EPI_ISL_720745\|20 | ···················· | ···················· | ···················· | ···················· | ···················· | ···················· |
| X | 728 | hCoV-19/England/MILK-BFF591/2020\|EPI_ISL_720746\|20 | ···················· | ···················· | ···················· | ···················· | ······T············· | ···················· |
| X | 729 | hCoV-19/England/MILK-BFF5FB/2020\|EPI_ISL_720747\|20 | ···················· | ···················· | ···················· | ···················· | ···················· | ···················· |
| X | 730 | hCoV-19/England/MILK-BFCD05/2020\|EPI_ISL_720748\|20 | ···················· | ···················· | ···················· | ···················· | ······T············· | ···················· |
| X | 731 | hCoV-19/England/ALDP-C04AD5/2020\|EPI_ISL_720750\|20 | ···················· | ···················· | ···················· | ···················· | ···················· | ···················· |
| X | 732 | hCoV-19/England/MILK-BFDEA7/2020\|EPI_ISL_720751\|20 | ···················· | ···················· | ···················· | ···················· | ···················· | ···················· |
| X | 733 | hCoV-19/England/QEUH-C01416/2020\|EPI_ISL_720752\|20 | ···················· | ···················· | ···················· | ···················· | ···················· | ···················· |
| X | 734 | hCoV-19/England/MILK-C00884/2020\|EPI_ISL_720753\|20 | ···················· | ···················· | ···················· | ···················· | ···················· | ···················· |
| X | 735 | hCoV-19/England/QEUH-C01434/2020\|EPI_ISL_720755\|20 | ···················· | ···················· | ···················· | ···················· | ···················· | ···················· |
| X | 736 | hCoV-19/England/ALDP-C04AF3/2020\|EPI_ISL_720756\|20 | ···················· | ···················· | ···················· | ···················· | ···················· | ···················· |
| X | 737 | hCoV-19/England/MILK-BFCD14/2020\|EPI_ISL_720760\|20 | ···················· | ···················· | ···················· | ···················· | ······T············· | ···················· |
| X | 738 | hCoV-19/England/QEUH-C03E61/2020\|EPI_ISL_720761\|20 | ···················· | ···················· | ···················· | ···················· | ···················· | ··A················· |
| X | 739 | hCoV-19/England/ALDP-C04EE8/2020\|EPI_ISL_720764\|20 | ···················· | ···················· | ···················· | ···················· | ···················· | ···················· |
| X | 740 | hCoV-19/England/QEUH-C03EE9/2020\|EPI_ISL_720767\|20 | ···················· | ···················· | ···················· | ···················· | ···················· | ···················· |
| X | 741 | hCoV-19/England/MILK-C006C6/2020\|EPI_ISL_720768\|20 | ···················· | ···················· | ···················· | ···················· | ···················· | ···················· |
| X | 742 | hCoV-19/England/MILK-BFD5EE/2020\|EPI_ISL_720769\|20 | ···················· | ···················· | ···················· | ···················· | ···················· | ···················· |
| X | 743 | hCoV-19/England/ALDP-C04A6C/2020\|EPI_ISL_720770\|20 | ···················· | ···················· | ···················· | ···················· | ···················· | ···················· |
| X | 744 | hCoV-19/England/MILK-BFD557/2020\|EPI_ISL_720771\|20 | ···················· | ···················· | ···················· | ···················· | ···················· | ···················· |
| X | 745 | hCoV-19/England/MILK-C0068A/2020\|EPI_ISL_720772\|20 | ···················· | ···················· | ···················· | ···················· | ···················· | ···················· |
| X | 746 | hCoV-19/England/ALDP-C05CB0/2020\|EPI_ISL_720773\|20 | ···················· | ···················· | ···················· | ···················· | ···················· | ···················· |
| X | 747 | hCoV-19/England/MILK-BFF968/2020\|EPI_ISL_720774\|20 | ···················· | ···················· | ···················· | ···················· | ···················· | ···················· |
| X | 748 | hCoV-19/England/MILK-C00893/2020\|EPI_ISL_720775\|20 | ···················· | ···················· | ···················· | ···················· | ···················· | ···················· |
| X | 749 | hCoV-19/England/QEUH-C01391/2020\|EPI_ISL_720776\|20 | ···················· | ···················· | ···················· | ···················· | ···················· | ···················· |
| X | 750 | hCoV-19/England/MILK-BFF573/2020\|EPI_ISL_720778\|20 | ···················· | ·A·················· | ···················· | ···················· | ···················· | ···················· |
| X | 751 | hCoV-19/England/ALDP-C04B0F/2020\|EPI_ISL_720779\|20 | ···················· | ···················· | ···················· | ···················· | ···················· | ···················· |
| X | 752 | hCoV-19/Wales/ALDP-C04914/2020\|EPI_ISL_720780\|2020 | ···················· | ···················· | ···················· | ···················· | ···················· | ···················· |
| X | 753 | hCoV-19/England/MILK-BFF5BF/2020\|EPI_ISL_720781\|20 | ···················· | ···················· | ···················· | ···················· | ···················· | ···················· |
| X | 754 | hCoV-19/England/MILK-C008DF/2020\|EPI_ISL_720782\|20 | ···················· | ···················· | ···················· | ···················· | ···················· | ···················· |
| X | 755 | hCoV-19/England/MILK-BFCE6C/2020\|EPI_ISL_720783\|20 | ···················· | ···················· | ···················· | ···················· | ···················· | ···················· |
| X | 756 | hCoV-19/England/MILK-BFCF87/2020\|EPI_ISL_720784\|20 | ···················· | ···················· | ···················· | ···················· | ···················· | ···················· |
| X | 757 | hCoV-19/England/MILK-BFF634/2020\|EPI_ISL_720785\|20 | ···················· | ·A·················· | ···················· | ···················· | ···················· | ···················· |
| X | 758 | hCoV-19/England/QEUH-C012A3/2020\|EPI_ISL_720787\|20 | ···················· | ···················· | ···················· | ···················· | ···················· | ···················· |
| X | 759 | hCoV-19/England/MILK-BFF6E9/2020\|EPI_ISL_720789\|20 | ···················· | ···················· | ···················· | ···················· | ···················· | ···················· |
| X | 760 | hCoV-19/England/MILK-BFF4A3/2020\|EPI_ISL_720791\|20 | ···················· | ···················· | ···················· | ···················· | ···················· | ···················· |
| X | 761 | hCoV-19/England/QEUH-C01355/2020\|EPI_ISL_720792\|20 | ···················· | ···················· | ···················· | ···················· | ···················· | ···················· |
| X | 762 | hCoV-19/England/MILK-BFCCCC/2020\|EPI_ISL_720794\|20 | ···················· | ···················· | ···················· | ···················· | ···················· | ···················· |
| X | 763 | hCoV-19/England/MILK-BFD593/2020\|EPI_ISL_720795\|20 | ···················· | ···················· | ···················· | ···················· | ···················· | ···················· |
| X | 764 | hCoV-19/England/ALDP-C04F12/2020\|EPI_ISL_720796\|20 | ···················· | ···················· | ···················· | ···················· | ···················· | ···················· |
| X | 765 | hCoV-19/England/MILK-BFF5EC/2020\|EPI_ISL_720797\|20 | ···················· | ···················· | ···················· | ···················· | ···················· | ···················· |
| X | 766 | hCoV-19/England/MILK-BFF6F8/2020\|EPI_ISL_720799\|20 | ···················· | ···················· | ···················· | ···················· | ···················· | ···················· |
| X | 767 | hCoV-19/England/EXET-13DA65/2020\|EPI_ISL_724274\|20 | ···················· | ···················· | ···················· | ···················· | ···················· | ···················· |
| X | 768 | hCoV-19/England/EXET-13DB08/2020\|EPI_ISL_724276\|20 | ···················· | ···················· | ···················· | ···················· | ···················· | ···················· |
| X | 769 | hCoV-19/Wales/QEUH-B2DE26/2020\|EPI_ISL_711001\|2020 | ···················· | ···················· | ···················· | ···················· | ···················· | ···················· |
| X | 770 | hCoV-19/Wales/QEUH-B2CA6F/2020\|EPI_ISL_711002\|2020 | ···················· | ···················· | ···················· | ···················· | ···················· | ···················· |
| X | 771 | hCoV-19/Wales/QEUH-B2B80C/2020\|EPI_ISL_711003\|2020 | ···················· | ···················· | ···················· | ···················· | ···················· | ···················· |
| X | 772 | hCoV-19/Wales/QEUH-B2B893/2020\|EPI_ISL_711004\|2020 | ···················· | ···················· | ···················· | ···················· | ···················· | ···················· |
| X | 773 | hCoV-19/Wales/QEUH-B2B74B/2020\|EPI_ISL_711005\|2020 | ···················· | ···················· | ···················· | ···················· | ···················· | ···················· |
| X | 774 | hCoV-19/Wales/QEUH-B2CA05/2020\|EPI_ISL_711006\|2020 | ···················· | ···················· | ···················· | ···················· | ···················· | ···················· |
| X | 775 | hCoV-19/Wales/QEUH-B2B365/2020\|EPI_ISL_711007\|2020 | ···················· | ···················· | ···················· | ···················· | ···················· | ···················· |
| X | 776 | hCoV-19/Wales/ALDP-B24724/2020\|EPI_ISL_711008\|2020 | ···················· | ···················· | ···················· | ···················· | ···················· | ···················· |
| X | 777 | hCoV-19/Wales/ALDP-B24C52/2020\|EPI_ISL_711009\|2020 | ···················· | ···················· | ···················· | ···················· | ···················· | ···················· |
| X | 778 | hCoV-19/Wales/ALDP-B24BBF/2020\|EPI_ISL_711010\|2020 | ···················· | ···················· | ···················· | ···················· | ···················· | ···················· |
| X | 779 | hCoV-19/Wales/ALDP-B24BCE/2020\|EPI_ISL_711012\|2020 | ···················· | ···················· | ···················· | ···················· | ···················· | ···················· |
| X | 780 | hCoV-19/Wales/ALDP-B24CBC/2020\|EPI_ISL_711013\|2020 | ···················· | ···················· | ···················· | ···················· | ···················· | ···················· |
| X | 781 | hCoV-19/Wales/QEUH-B2D99E/2020\|EPI_ISL_711014\|2020 | ···················· | ···················· | ···················· | ···················· | ···················· | ···················· |
| X | 782 | hCoV-19/Wales/ALDP-B24C70/2020\|EPI_ISL_711015\|2020 | ···················· | ···················· | ···················· | ···················· | ···················· | ···················· |
| X | 783 | hCoV-19/Wales/ALDP-B246DC/2020\|EPI_ISL_711016\|2020 | ···················· | ···················· | ···················· | ···················· | ···················· | ···················· |
| X | 784 | hCoV-19/Wales/ALDP-B246BE/2020\|EPI_ISL_711017\|2020 | ···················· | ···················· | ···················· | ···················· | ···················· | ···················· |
| X | 785 | hCoV-19/Wales/ALDP-B24BDD/2020\|EPI_ISL_711018\|2020 | ···················· | ···················· | ···················· | ···················· | ···················· | ···················· |
| X | 786 | hCoV-19/Wales/ALDP-B246EB/2020\|EPI_ISL_711019\|2020 | ···················· | ···················· | ···················· | ···················· | ···················· | ···················· |
| X | 787 | hCoV-19/Wales/ALDP-B24C61/2020\|EPI_ISL_711020\|2020 | ···················· | ···················· | ···················· | ···················· | ···················· | ···················· |
| X | 788 | hCoV-19/Wales/ALDP-B24C07/2020\|EPI_ISL_711021\|2020 | ···················· | ···················· | ···················· | ···················· | ···················· | ···················· |
| X | 789 | hCoV-19/Wales/ALDP-B24B91/2020\|EPI_ISL_711022\|2020 | ···················· | ···················· | ···················· | ···················· | ···················· | ···················· |
| X | 790 | hCoV-19/Wales/CAMC-B213BA/2020\|EPI_ISL_711024\|2020 | ···················· | ···················· | ···················· | ···················· | ···················· | ···················· |
| X | 791 | hCoV-19/Wales/CAMC-B22E30/2020\|EPI_ISL_711026\|2020 | ···················· | ···················· | ···················· | ···················· | ···················· | ···················· |
| X | 792 | hCoV-19/Wales/ALDP-B1A7E5/2020\|EPI_ISL_711027\|2020 | ···················· | ···················· | ···················· | ···················· | ···················· | ···················· |
| X | 793 | hCoV-19/Wales/ALDP-B1A83D/2020\|EPI_ISL_711028\|2020 | ···················· | ···················· | ···················· | ···················· | ···················· | ···················· |
| X | 794 | hCoV-19/Wales/ALDP-B1AB25/2020\|EPI_ISL_711029\|2020 | ···················· | ···················· | ···················· | ···················· | ···················· | ···················· |
| X | 795 | hCoV-19/Wales/ALDP-B1A8C4/2020\|EPI_ISL_711030\|2020 | ···················· | ···················· | ···················· | ···················· | ···················· | ···················· |
| X | 796 | hCoV-19/Wales/ALDP-B1A8D3/2020\|EPI_ISL_711031\|2020 | ···················· | ···················· | ···················· | ···················· | ··A················· | ···················· |
| X | 797 | hCoV-19/Wales/ALDP-B1AB16/2020\|EPI_ISL_711032\|2020 | ···················· | ···················· | ···················· | ···················· | ···················· | ···················· |
| X | 798 | hCoV-19/Wales/ALDP-B1A82E/2020\|EPI_ISL_711033\|2020 | ···················· | ···················· | ···················· | ···················· | ···················· | ···················· |
| X | 799 | hCoV-19/Wales/ALDP-B19F2A/2020\|EPI_ISL_711034\|2020 | ···················· | ···················· | ···················· | ···················· | ···················· | ···················· |
| X | 800 | hCoV-19/Wales/ALDP-B1AAFB/2020\|EPI_ISL_711036\|2020 | ···················· | ···················· | ···················· | ···················· | ···················· | ···················· |
| X | 801 | hCoV-19/Wales/ALDP-B1A053/2020\|EPI_ISL_711037\|2020 | ···················· | ···················· | ···················· | ···················· | ···················· | ···················· |
| X | 802 | hCoV-19/Wales/CAMC-B20BD1/2020\|EPI_ISL_711038\|2020 | ···················· | ···················· | ···················· | ···················· | ···················· | ···················· |
| X | 803 | hCoV-19/Wales/ALDP-B19FA2/2020\|EPI_ISL_711039\|2020 | ···················· | ···················· | ···················· | ···················· | ···················· | ···················· |
| X | 804 | hCoV-19/Wales/ALDP-B1A563/2020\|EPI_ISL_711040\|2020 | ···················· | ···················· | ···················· | ···················· | ···················· | ···················· |
| X | 805 | hCoV-19/Wales/ALDP-B1A581/2020\|EPI_ISL_711041\|2020 | ···················· | ···················· | ···················· | ···················· | ···················· | ···················· |
| X | 806 | hCoV-19/Wales/ALDP-B1A90D/2020\|EPI_ISL_711042\|2020 | ···················· | ···················· | ···················· | ···················· | ···················· | ···················· |

| | | | | | | | |
|---|---|---|---|---|---|---|---|
| X | 807 hCoV-19/Wales/ALDP-B19F75/2020\|EPI_ISL_711045\|2020 | ···················· | ···················· | ···················· | ···················· | ···················· | ···················· |
| X | 808 hCoV-19/Wales/ALDP-B1AB8F/2020\|EPI_ISL_711047\|2020 | ···················· | ···················· | ···················· | ···················· | ···················· | ···················· |
| X | 809 hCoV-19/Wales/QEUH-AF076E/2020\|EPI_ISL_711049\|2020 | ···················· | ···················· | ···················· | ···················· | ···················· | ···················· |
| X | 810 hCoV-19/Wales/QEUH-AF09B3/2020\|EPI_ISL_711052\|2020 | ···················· | ···················· | ···················· | ···················· | ···················· | ···················· |
| X | 811 hCoV-19/Wales/QEUH-AF0643/2020\|EPI_ISL_711053\|2020 | ···················· | ···················· | ···················· | ···················· | ···················· | ···················· |
| X | 812 hCoV-19/Wales/QEUH-AF0C41/2020\|EPI_ISL_711054\|2020 | ···················· | ···················· | ···················· | ···················· | ···················· | ···················· |
| X | 813 hCoV-19/Tunisia/ADAGE-18503/2020\|EPI_ISL_711057\|20 | ···················· | ···················· | ···················· | ···················· | ···················· | ···················· |
| X | 814 hCoV-19/Denmark/DCGC-13792/2020\|EPI_ISL_712300\|202 | ···················· | ···················· | ···················· | ···················· | ···················· | ···················· |
| X | 815 hCoV-19/Denmark/DCGC-13107/2020\|EPI_ISL_712301\|202 | ···················· | ···················· | ···················· | ···················· | ···················· | ···················· |
| X | 816 hCoV-19/Denmark/DCGC-13310/2020\|EPI_ISL_712302\|202 | ···················· | ···················· | ···················· | ···················· | ···················· | ···················· |
| X | 817 hCoV-19/Denmark/DCGC-13556/2020\|EPI_ISL_712303\|202 | ···················· | ···················· | ···················· | ···················· | ···················· | ···················· |
| X | 818 hCoV-19/Denmark/DCGC-13202/2020\|EPI_ISL_712304\|202 | ···················· | ···················· | ···················· | ···················· | ···················· | ···················· |
| X | 819 hCoV-19/Denmark/DCGC-12920/2020\|EPI_ISL_712321\|202 | ···················· | ···················· | ···················· | ···················· | ···················· | ···················· |
| X | 820 hCoV-19/Denmark/DCGC-13097/2020\|EPI_ISL_712322\|202 | ···················· | ···················· | ···················· | ···················· | ···················· | ···················· |
| X | 821 hCoV-19/Denmark/DCGC-13131/2020\|EPI_ISL_712323\|202 | ···················· | ···················· | ···················· | ···················· | ···················· | ···················· |
| X | 822 hCoV-19/Denmark/DCGC-13178/2020\|EPI_ISL_712324\|202 | ···················· | ···················· | ···················· | ···················· | ···················· | ···················· |
| X | 823 hCoV-19/Denmark/DCGC-13182/2020\|EPI_ISL_712325\|202 | ···················· | ···················· | ···················· | ···················· | ···················· | ···················· |
| X | 824 hCoV-19/Denmark/DCGC-13306/2020\|EPI_ISL_712326\|202 | ···················· | ···················· | ···················· | ···················· | ···················· | ···················· |
| X | 825 hCoV-19/Denmark/DCGC-13468/2020\|EPI_ISL_712327\|202 | ···················· | ···················· | ···················· | ···················· | ···················· | ···················· |
| X | 826 hCoV-19/Denmark/DCGC-13540/2020\|EPI_ISL_712328\|202 | ···················· | ···················· | ···················· | ···················· | ···················· | ···················· |
| X | 827 hCoV-19/Denmark/DCGC-13643/2020\|EPI_ISL_712329\|202 | ···················· | ···················· | ···················· | ···················· | ···················· | ···················· |
| X | 828 hCoV-19/Denmark/DCGC-13668/2020\|EPI_ISL_712330\|202 | ···················· | ···················· | ···················· | ···················· | ···················· | ···················· |
| X | 829 hCoV-19/Denmark/DCGC-13702/2020\|EPI_ISL_712331\|202 | ···················· | ···················· | ···················· | ···················· | ···················· | ···················· |
| X | 830 hCoV-19/Denmark/DCGC-13738/2020\|EPI_ISL_712332\|202 | ···················· | ···················· | ···················· | ···················· | ···················· | ···················· |
| X | 831 hCoV-19/Denmark/DCGC-13743/2020\|EPI_ISL_712333\|202 | ···················· | ···················· | ···················· | ···················· | ···················· | ···················· |
| X | 832 hCoV-19/Denmark/DCGC-13762/2020\|EPI_ISL_712334\|202 | ···················· | ···················· | ···················· | ···················· | ···················· | ···················· |
| X | 833 hCoV-19/Denmark/DCGC-13882/2020\|EPI_ISL_712335\|202 | ···················· | ···················· | ···················· | ···················· | ···················· | ···················· |
| X | 834 hCoV-19/Denmark/DCGC-13904/2020\|EPI_ISL_712336\|202 | ···················· | ···················· | ···················· | ···················· | ···················· | ···················· |
| X | 835 hCoV-19/Denmark/DCGC-13905/2020\|EPI_ISL_712337\|202 | ···················· | ···················· | ···················· | ···················· | ···················· | ···················· |
| X | 836 hCoV-19/Denmark/DCGC-12928/2020\|EPI_ISL_712340\|202 | ···················· | ···················· | ···················· | ···················· | ···················· | ···················· |
| X | 837 hCoV-19/Denmark/DCGC-12944/2020\|EPI_ISL_712341\|202 | ···················· | ···················· | ···················· | ···················· | ···················· | ···················· |
| X | 838 hCoV-19/Denmark/DCGC-12952/2020\|EPI_ISL_712342\|202 | ···················· | ···················· | ···················· | ···················· | ···················· | ···················· |
| X | 839 hCoV-19/Denmark/DCGC-12960/2020\|EPI_ISL_712343\|202 | ···················· | ···················· | ···················· | ···················· | ···················· | ···················· |
| X | 840 hCoV-19/Denmark/DCGC-12970/2020\|EPI_ISL_712344\|202 | ···················· | ···················· | ···················· | ···················· | ···················· | ···················· |
| X | 841 hCoV-19/Denmark/DCGC-12976/2020\|EPI_ISL_712345\|202 | ···················· | ···················· | ···················· | ···················· | ···················· | ···················· |
| X | 842 hCoV-19/Denmark/DCGC-12990/2020\|EPI_ISL_712346\|202 | ···················· | ···················· | ···················· | ···················· | ···················· | ···················· |
| X | 843 hCoV-19/Denmark/DCGC-13022/2020\|EPI_ISL_712347\|202 | ···················· | ···················· | ···················· | ···················· | ···················· | ···················· |
| X | 844 hCoV-19/Denmark/DCGC-13025/2020\|EPI_ISL_712348\|202 | ···················· | ···················· | ···················· | ···················· | ···················· | ···················· |
| X | 845 hCoV-19/Denmark/DCGC-13051/2020\|EPI_ISL_712349\|202 | ···················· | ···················· | ···················· | ···················· | ···················· | ···················· |
| X | 846 hCoV-19/Denmark/DCGC-13057/2020\|EPI_ISL_712350\|202 | ···················· | ···················· | ···················· | ···················· | ···················· | ···················· |
| X | 847 hCoV-19/Denmark/DCGC-13071/2020\|EPI_ISL_712351\|202 | ···················· | ···················· | ···················· | ···················· | ···················· | ···················· |
| X | 848 hCoV-19/Denmark/DCGC-13074/2020\|EPI_ISL_712352\|202 | ···················· | ···················· | ···················· | ···················· | ··C················· | ···················· |
| X | 849 hCoV-19/Denmark/DCGC-13085/2020\|EPI_ISL_712353\|202 | ···················· | ···················· | ···················· | ···················· | ···················· | ···················· |
| X | 850 hCoV-19/Denmark/DCGC-13089/2020\|EPI_ISL_712354\|202 | ···················· | ···················· | ···················· | ···················· | ···················· | ···················· |
| X | 851 hCoV-19/Denmark/DCGC-13090/2020\|EPI_ISL_712355\|202 | ···················· | ···················· | ···················· | ···················· | ···················· | ···················· |
| X | 852 hCoV-19/Denmark/DCGC-13092/2020\|EPI_ISL_712356\|202 | ···················· | ···················· | ···················· | ···················· | ···················· | ···················· |
| X | 853 hCoV-19/Denmark/DCGC-13101/2020\|EPI_ISL_712357\|202 | ···················· | ···················· | ···················· | ···················· | ···················· | ···················· |
| X | 854 hCoV-19/Denmark/DCGC-13114/2020\|EPI_ISL_712358\|202 | ···················· | ···················· | ···················· | ···················· | ···················· | ···················· |
| X | 855 hCoV-19/Denmark/DCGC-13134/2020\|EPI_ISL_712359\|202 | ···················· | ···················· | ···················· | ···················· | ···················· | ···················· |
| X | 856 hCoV-19/Denmark/DCGC-13169/2020\|EPI_ISL_712360\|202 | ···················· | ···················· | ···················· | ···················· | ···················· | ···················· |
| X | 857 hCoV-19/Denmark/DCGC-13179/2020\|EPI_ISL_712361\|202 | ···················· | ···················· | ···················· | ···················· | ···················· | ···················· |
| X | 858 hCoV-19/Denmark/DCGC-13191/2020\|EPI_ISL_712362\|202 | ···················· | ···················· | ···················· | ···················· | ···················· | ···················· |
| X | 859 hCoV-19/Denmark/DCGC-13197/2020\|EPI_ISL_712363\|202 | ···················· | ···················· | ···················· | ···················· | ···················· | ···················· |
| X | 860 hCoV-19/Denmark/DCGC-13223/2020\|EPI_ISL_712364\|202 | ···················· | ···················· | ···················· | ···················· | ···················· | ···················· |
| X | 861 hCoV-19/Denmark/DCGC-13226/2020\|EPI_ISL_712365\|202 | ···················· | ···················· | ···················· | ···················· | ···················· | ···················· |
| X | 862 hCoV-19/Denmark/DCGC-13227/2020\|EPI_ISL_712366\|202 | ···················· | ···················· | ···················· | ···················· | ···················· | ···················· |
| X | 863 hCoV-19/Denmark/DCGC-13228/2020\|EPI_ISL_712367\|202 | ···················· | ···················· | ···················· | ···················· | ···················· | ···················· |
| X | 864 hCoV-19/Denmark/DCGC-13235/2020\|EPI_ISL_712368\|202 | ···················· | ···················· | ···················· | ···················· | ···················· | ···················· |
| X | 865 hCoV-19/Denmark/DCGC-13251/2020\|EPI_ISL_712369\|202 | ···················· | ···················· | ···················· | ···················· | ···················· | ···················· |
| X | 866 hCoV-19/Denmark/DCGC-13298/2020\|EPI_ISL_712370\|202 | ···················· | ···················· | ···················· | ···················· | ···················· | ···················· |
| X | 867 hCoV-19/Denmark/DCGC-13319/2020\|EPI_ISL_712371\|202 | ···················· | ···················· | ···················· | ···················· | ···················· | ···················· |
| X | 868 hCoV-19/Denmark/DCGC-13321/2020\|EPI_ISL_712372\|202 | ···················· | ···················· | ···················· | ···················· | ···················· | ···················· |
| X | 869 hCoV-19/Denmark/DCGC-13346/2020\|EPI_ISL_712373\|202 | ···················· | ···················· | ···················· | ···················· | ···················· | ···················· |
| X | 870 hCoV-19/Denmark/DCGC-13348/2020\|EPI_ISL_712374\|202 | ···················· | ···················· | ···················· | ···················· | ···················· | ···················· |
| X | 871 hCoV-19/Denmark/DCGC-13352/2020\|EPI_ISL_712375\|202 | ···················· | ···················· | ···················· | ···················· | ···················· | ···················· |
| X | 872 hCoV-19/Denmark/DCGC-13354/2020\|EPI_ISL_712376\|202 | ···················· | ···················· | ···················· | ···················· | ···················· | ···················· |
| X | 873 hCoV-19/Denmark/DCGC-13356/2020\|EPI_ISL_712377\|202 | ···················· | ···················· | ···················· | ···················· | ···················· | ···················· |
| X | 874 hCoV-19/Denmark/DCGC-13360/2020\|EPI_ISL_712378\|202 | ···················· | ···················· | ···················· | ···················· | ···················· | ···················· |
| X | 875 hCoV-19/Denmark/DCGC-13374/2020\|EPI_ISL_712379\|202 | ···················· | ···················· | ···················· | ···················· | ···················· | ···················· |
| X | 876 hCoV-19/Denmark/DCGC-13397/2020\|EPI_ISL_712380\|202 | ···················· | ···················· | ···················· | ···················· | ···················· | ···················· |
| X | 877 hCoV-19/Denmark/DCGC-13403/2020\|EPI_ISL_712381\|202 | ···················· | ···················· | ···················· | ···················· | ···················· | ···················· |
| X | 878 hCoV-19/Denmark/DCGC-13421/2020\|EPI_ISL_712382\|202 | ···················· | ···················· | ···················· | ···················· | ···················· | ···················· |
| X | 879 hCoV-19/Denmark/DCGC-13422/2020\|EPI_ISL_712383\|202 | ···················· | ···················· | ···················· | ···················· | ···················· | ···················· |
| X | 880 hCoV-19/Denmark/DCGC-13424/2020\|EPI_ISL_712384\|202 | ···················· | ···················· | ···················· | ···················· | ···················· | ···················· |
| X | 881 hCoV-19/Denmark/DCGC-13426/2020\|EPI_ISL_712385\|202 | ···················· | ···················· | ···················· | ···················· | ···················· | ···················· |
| X | 882 hCoV-19/Denmark/DCGC-13444/2020\|EPI_ISL_712386\|202 | ···················· | ···················· | ···················· | ···················· | ···················· | ···················· |
| X | 883 hCoV-19/Denmark/DCGC-13465/2020\|EPI_ISL_712387\|202 | ···················· | ···················· | ···················· | ···················· | ···················· | ···················· |
| X | 884 hCoV-19/Denmark/DCGC-13472/2020\|EPI_ISL_712388\|202 | ···················· | ···················· | ···················· | ···················· | ··C················· | ···················· |
| X | 885 hCoV-19/Denmark/DCGC-13484/2020\|EPI_ISL_712389\|202 | ···················· | ···················· | ···················· | ···················· | ···················· | ···················· |
| X | 886 hCoV-19/Denmark/DCGC-13516/2020\|EPI_ISL_712390\|202 | ···················· | ···················· | ···················· | ···················· | ···················· | ···················· |

| | | | | | | | |
|---|---|---|---|---|---|---|---|
| X | 887 hCoV-19/Denmark/DCGC-13521/2020\|EPI_ISL_712391\|202 | ···················· | ···················· | ···················· | ···················· | ···················· | ···················· |
| X | 888 hCoV-19/Denmark/DCGC-13557/2020\|EPI_ISL_712392\|202 | ···················· | ···················· | ···················· | ···················· | ···················· | ···················· |
| X | 889 hCoV-19/Denmark/DCGC-13565/2020\|EPI_ISL_712393\|202 | ···················· | ···················· | ···················· | ···················· | ··C················· | ···················· |
| X | 890 hCoV-19/Denmark/DCGC-13582/2020\|EPI_ISL_712394\|202 | ···················· | ···················· | ···················· | ···················· | ···················· | ···················· |
| X | 891 hCoV-19/Denmark/DCGC-13596/2020\|EPI_ISL_712395\|202 | ···················· | ···················· | ···················· | ···················· | ···················· | ···················· |
| X | 892 hCoV-19/Denmark/DCGC-13613/2020\|EPI_ISL_712396\|202 | ···················· | ···················· | ···················· | ···················· | ···T················ | ···················· |
| X | 893 hCoV-19/Denmark/DCGC-13664/2020\|EPI_ISL_712397\|202 | ···················· | ···················· | ···················· | ···················· | ···················· | ···················· |
| X | 894 hCoV-19/Denmark/DCGC-13690/2020\|EPI_ISL_712398\|202 | ···················· | ···················· | ···················· | ···················· | ···················· | ···················· |
| X | 895 hCoV-19/Denmark/DCGC-13694/2020\|EPI_ISL_712399\|202 | ···················· | ···················· | ···················· | ···················· | ···················· | ···················· |
| X | 896 hCoV-19/Denmark/DCGC-15069/2020\|EPI_ISL_713616\|202 | ···················· | ···················· | ···················· | ···················· | ···················· | ···················· |
| X | 897 hCoV-19/Denmark/DCGC-15070/2020\|EPI_ISL_713617\|202 | ···················· | ···················· | ···················· | ···················· | ···················· | ···················· |
| X | 898 hCoV-19/Denmark/DCGC-15071/2020\|EPI_ISL_713618\|202 | ···················· | ···················· | ···················· | ···················· | ···················· | ···················· |
| X | 899 hCoV-19/Denmark/DCGC-15073/2020\|EPI_ISL_713619\|202 | ···················· | ···················· | ···················· | ···················· | ···················· | ···················· |
| X | 900 hCoV-19/Denmark/DCGC-15074/2020\|EPI_ISL_713620\|202 | ···················· | ···················· | ···················· | ···················· | ···················· | ···················· |
| X | 901 hCoV-19/Denmark/DCGC-15075/2020\|EPI_ISL_713621\|202 | ···················· | ···················· | ···················· | ···················· | ···················· | ···················· |
| X | 902 hCoV-19/Denmark/DCGC-15076/2020\|EPI_ISL_713622\|202 | ···················· | ···················· | ···················· | ···················· | ···················· | ···················· |
| X | 903 hCoV-19/Denmark/DCGC-15080/2020\|EPI_ISL_713623\|202 | ···················· | ···················· | ···················· | ···················· | ···················· | ···················· |
| X | 904 hCoV-19/Denmark/DCGC-15083/2020\|EPI_ISL_713624\|202 | ···················· | ···················· | ···················· | ···················· | ···················· | ···················· |
| X | 905 hCoV-19/Denmark/DCGC-15084/2020\|EPI_ISL_713625\|202 | ···················· | ···················· | ···················· | ···················· | ···················· | ···················· |
| X | 906 hCoV-19/Denmark/DCGC-15094/2020\|EPI_ISL_713626\|202 | ···················· | ···A················ | ···················· | ···················· | ···················· | ···················· |
| X | 907 hCoV-19/Denmark/DCGC-15095/2020\|EPI_ISL_713627\|202 | ···················· | ···················· | ···················· | ···················· | ···················· | ···················· |
| X | 908 hCoV-19/Denmark/DCGC-15096/2020\|EPI_ISL_713628\|202 | ···················· | ···················· | ···················· | ···················· | ···················· | ···················· |
| X | 909 hCoV-19/Denmark/DCGC-15099/2020\|EPI_ISL_713629\|202 | ···················· | ···A················ | ···················· | ···················· | ···················· | ···················· |
| X | 910 hCoV-19/Denmark/DCGC-15100/2020\|EPI_ISL_713630\|202 | ···················· | ···················· | ···················· | ···················· | ···················· | ···················· |
| X | 911 hCoV-19/Denmark/DCGC-15101/2020\|EPI_ISL_713631\|202 | ···················· | ···················· | ···················· | ···················· | ···················· | ···················· |
| X | 912 hCoV-19/Denmark/DCGC-15102/2020\|EPI_ISL_713632\|202 | ···················· | ·A·················· | ···················· | ···················· | ···················· | ··A················· |
| X | 913 hCoV-19/Denmark/DCGC-15106/2020\|EPI_ISL_713633\|202 | ···················· | ···················· | ···················· | ···················· | ···················· | ···················· |
| X | 914 hCoV-19/Denmark/DCGC-15107/2020\|EPI_ISL_713634\|202 | ···················· | ···················· | ···················· | ···················· | ···················· | ···················· |
| X | 915 hCoV-19/Denmark/DCGC-15109/2020\|EPI_ISL_713635\|202 | ···················· | ···················· | ···················· | ···················· | ···················· | ···················· |
| X | 916 hCoV-19/Denmark/DCGC-15111/2020\|EPI_ISL_713636\|202 | ···················· | ···················· | ···················· | ···················· | ···················· | ···················· |
| X | 917 hCoV-19/Denmark/DCGC-15112/2020\|EPI_ISL_713637\|202 | ···················· | ···················· | ···················· | ···················· | ···················· | ···················· |
| X | 918 hCoV-19/Denmark/DCGC-15115/2020\|EPI_ISL_713640\|202 | ···················· | ···················· | ···················· | ···················· | ···················· | ···················· |
| X | 919 hCoV-19/Denmark/DCGC-15119/2020\|EPI_ISL_713641\|202 | ···················· | ···················· | ···················· | ···················· | ···················· | ···················· |
| X | 920 hCoV-19/Denmark/DCGC-15124/2020\|EPI_ISL_713642\|202 | ···················· | ·A·················· | ···················· | ···················· | ···················· | ··A················· |
| X | 921 hCoV-19/Denmark/DCGC-15125/2020\|EPI_ISL_713643\|202 | ···················· | ···················· | ···················· | ···················· | ···················· | ···················· |
| X | 922 hCoV-19/Denmark/DCGC-15127/2020\|EPI_ISL_713644\|202 | ···················· | ···················· | ···················· | ···················· | ···················· | ···················· |
| X | 923 hCoV-19/Denmark/DCGC-15133/2020\|EPI_ISL_713649\|202 | ···················· | ···················· | ···················· | ···················· | ···················· | ···················· |
| X | 924 hCoV-19/Denmark/DCGC-15134/2020\|EPI_ISL_713650\|202 | ···················· | ···················· | ···················· | ···················· | ···················· | ···················· |
| X | 925 hCoV-19/Denmark/DCGC-15136/2020\|EPI_ISL_713651\|202 | ···················· | ···················· | ···················· | ···················· | ···················· | ···················· |
| X | 926 hCoV-19/Denmark/DCGC-15137/2020\|EPI_ISL_713652\|202 | ···················· | ···················· | ···················· | ···················· | ···················· | ···················· |
| X | 927 hCoV-19/Denmark/DCGC-15140/2020\|EPI_ISL_713653\|202 | ···················· | ···················· | ···················· | ···················· | ···················· | ···················· |
| X | 928 hCoV-19/Denmark/DCGC-15141/2020\|EPI_ISL_713654\|202 | ···················· | ···················· | ···················· | ···················· | ···················· | ···················· |
| X | 929 hCoV-19/Denmark/DCGC-15142/2020\|EPI_ISL_713655\|202 | ···················· | ···A················ | ···················· | ···················· | ···················· | ···················· |
| X | 930 hCoV-19/Denmark/DCGC-15143/2020\|EPI_ISL_713656\|202 | ···················· | ···················· | ···················· | ···················· | ···················· | ···················· |
| X | 931 hCoV-19/Denmark/DCGC-15145/2020\|EPI_ISL_713657\|202 | ···················· | ···················· | ···················· | ···················· | ···················· | ···················· |
| X | 932 hCoV-19/Denmark/DCGC-15146/2020\|EPI_ISL_713658\|202 | ···················· | ···················· | ···················· | ···················· | ···················· | ···················· |
| X | 933 hCoV-19/Denmark/DCGC-15147/2020\|EPI_ISL_713659\|202 | ···················· | ···················· | ···················· | ···················· | ···················· | ···················· |
| X | 934 hCoV-19/Denmark/DCGC-15149/2020\|EPI_ISL_713660\|202 | ···················· | ···················· | ···················· | ···················· | ···················· | ···················· |
| X | 935 hCoV-19/Denmark/DCGC-15150/2020\|EPI_ISL_713661\|202 | ···················· | ···················· | ···················· | ···················· | ···················· | ···················· |
| X | 936 hCoV-19/Denmark/DCGC-15151/2020\|EPI_ISL_713662\|202 | ···················· | ···················· | ···················· | ···················· | ···················· | ···················· |
| X | 937 hCoV-19/Denmark/DCGC-15153/2020\|EPI_ISL_713663\|202 | ···················· | ···················· | ···················· | ···················· | ···················· | ···················· |
| X | 938 hCoV-19/Denmark/DCGC-15156/2020\|EPI_ISL_713664\|202 | ···················· | ···················· | ···················· | ···················· | ···················· | ···················· |
| X | 939 hCoV-19/Denmark/DCGC-15157/2020\|EPI_ISL_713665\|202 | ···················· | ···················· | ···················· | ···················· | ···················· | ···················· |
| X | 940 hCoV-19/Denmark/DCGC-15158/2020\|EPI_ISL_713666\|202 | ···················· | ···················· | ···················· | ···················· | ···················· | ···················· |
| X | 941 hCoV-19/Denmark/DCGC-15161/2020\|EPI_ISL_713667\|202 | ···················· | ···················· | ···················· | ···················· | ···················· | ···················· |
| X | 942 hCoV-19/Denmark/DCGC-15162/2020\|EPI_ISL_713668\|202 | ···················· | ···················· | ···················· | ···················· | ···················· | ···················· |
| X | 943 hCoV-19/Denmark/DCGC-15164/2020\|EPI_ISL_713669\|202 | ···················· | ···················· | ···················· | ···················· | ·····T·············· | ···················· |
| X | 944 hCoV-19/Denmark/DCGC-15165/2020\|EPI_ISL_713670\|202 | ···················· | ···················· | ···················· | ···················· | ···················· | ···················· |
| X | 945 hCoV-19/Denmark/DCGC-15169/2020\|EPI_ISL_713671\|202 | ···················· | ···················· | ···················· | ···················· | ···················· | ···················· |
| X | 946 hCoV-19/Denmark/DCGC-15170/2020\|EPI_ISL_713672\|202 | ···················· | ···················· | ···················· | ···················· | ···················· | ···················· |
| X | 947 hCoV-19/Denmark/DCGC-15173/2020\|EPI_ISL_713673\|202 | ···················· | ···················· | ···················· | ···················· | ···················· | ···················· |
| X | 948 hCoV-19/Denmark/DCGC-15174/2020\|EPI_ISL_713674\|202 | ···················· | ···················· | ···················· | ···················· | ·····T·············· | ···················· |
| X | 949 hCoV-19/Denmark/DCGC-15175/2020\|EPI_ISL_713675\|202 | ···················· | ···················· | ···················· | ···················· | ···················· | ···················· |
| X | 950 hCoV-19/Denmark/DCGC-15176/2020\|EPI_ISL_713676\|202 | ···················· | ·A·················· | ···················· | ···················· | ···················· | ··A················· |
| X | 951 hCoV-19/Denmark/DCGC-15178/2020\|EPI_ISL_713677\|202 | ···················· | ···················· | ···················· | ···················· | ···················· | ···················· |
| X | 952 hCoV-19/Denmark/DCGC-15181/2020\|EPI_ISL_713678\|202 | ···················· | ···················· | ···················· | ···················· | ···················· | ···················· |
| X | 953 hCoV-19/Denmark/DCGC-15182/2020\|EPI_ISL_713679\|202 | ···················· | ···················· | ···················· | ···················· | ···················· | ···················· |
| X | 954 hCoV-19/Denmark/DCGC-15184/2020\|EPI_ISL_713680\|202 | ···················· | ·A·················· | ···················· | ···················· | ···················· | ··A················· |
| X | 955 hCoV-19/Denmark/DCGC-15185/2020\|EPI_ISL_713681\|202 | ···················· | ···················· | ···················· | ···················· | ···················· | ···················· |
| X | 956 hCoV-19/Denmark/DCGC-15187/2020\|EPI_ISL_713682\|202 | ···················· | ···················· | ···················· | ···················· | ···················· | ···················· |
| X | 957 hCoV-19/Denmark/DCGC-15189/2020\|EPI_ISL_713683\|202 | ···················· | ···················· | ···················· | ···················· | ···················· | ···················· |
| X | 958 hCoV-19/Denmark/DCGC-15191/2020\|EPI_ISL_713684\|202 | ···················· | ···················· | ···················· | ···················· | ···················· | ···················· |
| X | 959 hCoV-19/Denmark/DCGC-15193/2020\|EPI_ISL_713685\|202 | ···················· | ···················· | ···················· | ···················· | ···················· | ···················· |
| X | 960 hCoV-19/Denmark/DCGC-15196/2020\|EPI_ISL_713686\|202 | ···················· | ···················· | ···················· | ···················· | ···················· | ···················· |
| X | 961 hCoV-19/Denmark/DCGC-15198/2020\|EPI_ISL_713687\|202 | ···················· | ···················· | ···················· | ···················· | ···················· | ···················· |
| X | 962 hCoV-19/Denmark/DCGC-15199/2020\|EPI_ISL_713688\|202 | ···················· | ···················· | ···················· | ···················· | ···················· | ···················· |
| X | 963 hCoV-19/Denmark/DCGC-15200/2020\|EPI_ISL_713689\|202 | ···················· | ···················· | ···················· | ···················· | ···················· | ···················· |
| X | 964 hCoV-19/Denmark/DCGC-15201/2020\|EPI_ISL_713690\|202 | ···················· | ···················· | ···················· | ···················· | ···················· | ···················· |
| X | 965 hCoV-19/Denmark/DCGC-15202/2020\|EPI_ISL_713691\|202 | ···················· | ···················· | ···················· | ···················· | ···················· | ···················· |
| X | 966 hCoV-19/Denmark/DCGC-15205/2020\|EPI_ISL_713692\|202 | ···················· | ···················· | ···················· | ···················· | ···················· | ···················· |

| | | | | | | | | |
|---|---|---|---|---|---|---|---|---|
| X | 967 | hCoV-19/Denmark/DCGC-15207/2020\|EPI_ISL_713693\|202 | ···················· | ···················· | ···················· | ···················· | ···················· | ···················· |
| X | 968 | hCoV-19/Denmark/DCGC-15208/2020\|EPI_ISL_713694\|202 | ···················· | ···················· | ···················· | ···················· | ···················· | ···················· |
| X | 969 | hCoV-19/Denmark/DCGC-15210/2020\|EPI_ISL_713695\|202 | ···················· | ···················· | ···················· | ···················· | ···················· | ···················· |
| X | 970 | hCoV-19/Denmark/DCGC-15211/2020\|EPI_ISL_713696\|202 | ···················· | ·A·················· | ···················· | ···················· | ···················· | ···A················ |
| X | 971 | hCoV-19/Denmark/DCGC-15212/2020\|EPI_ISL_713697\|202 | ···················· | ···················· | ···················· | ···················· | ···················· | ···················· |
| X | 972 | hCoV-19/Denmark/DCGC-15214/2020\|EPI_ISL_713698\|202 | ···················· | ···················· | ···················· | ···················· | ···················· | ···················· |
| X | 973 | hCoV-19/Denmark/DCGC-15215/2020\|EPI_ISL_713699\|202 | ···················· | ···················· | ···················· | ···················· | ···················· | ···················· |
| X | 974 | hCoV-19/England/204861303/2020\|EPI_ISL_723016\|2020 | ···················· | ···················· | ···················· | ···················· | ···················· | ···················· |
| X | 975 | hCoV-19/England/204880322/2020\|EPI_ISL_723017\|2020 | ···················· | ···················· | ···················· | ···················· | ···················· | ···················· |
| X | 976 | hCoV-19/England/204891341/2020\|EPI_ISL_723018\|2020 | ···················· | ···················· | ···················· | ···················· | ···················· | ···················· |
| X | 977 | hCoV-19/England/204891344/2020\|EPI_ISL_723019\|2020 | ···················· | ···················· | ···················· | ···················· | ···················· | ···················· |
| X | 978 | hCoV-19/England/204891537/2020\|EPI_ISL_723020\|2020 | ···················· | ···················· | ···················· | ···················· | ···················· | ···················· |
| X | 979 | hCoV-19/England/204900599/2020\|EPI_ISL_723021\|2020 | ···················· | ···················· | ···················· | ···················· | ···················· | ···················· |
| X | 980 | hCoV-19/England/204900620/2020\|EPI_ISL_723022\|2020 | ···················· | ···················· | ···················· | ···················· | ···················· | ···················· |
| X | 981 | hCoV-19/England/204922366/2020\|EPI_ISL_723023\|2020 | ···················· | ···················· | ···················· | ···················· | ···················· | ···················· |
| X | 982 | hCoV-19/England/204960161/2020\|EPI_ISL_723024\|2020 | ···················· | ···················· | ···················· | ···················· | ···················· | ···················· |
| X | 983 | hCoV-19/England/204980132/2020\|EPI_ISL_723025\|2020 | ···················· | ·A·················· | ···················· | ···················· | ···················· | ···A················ |
| X | 984 | hCoV-19/England/204980135/2020\|EPI_ISL_723026\|2020 | ···················· | ·A·················· | ···················· | ···················· | ···················· | ···A················ |
| X | 985 | hCoV-19/England/204980142/2020\|EPI_ISL_723027\|2020 | ···················· | ·A·················· | ···················· | ···················· | ···················· | ···A················ |
| X | 986 | hCoV-19/England/204980318/2020\|EPI_ISL_723028\|2020 | ···················· | ···················· | ···················· | ···················· | ···················· | ···················· |
| X | 987 | hCoV-19/England/204980327/2020\|EPI_ISL_723029\|2020 | ···················· | ···················· | ···················· | ···················· | ···················· | ···················· |
| X | 988 | hCoV-19/England/204981804/2020\|EPI_ISL_723030\|2020 | ···················· | ···················· | ···················· | ···················· | ···················· | ···················· |
| X | 989 | hCoV-19/England/204990348/2020\|EPI_ISL_723031\|2020 | ···················· | ·A·················· | ···················· | ···················· | ···················· | ···A················ |
| X | 990 | hCoV-19/England/204991061/2020\|EPI_ISL_723032\|2020 | ···················· | ···················· | ···················· | ···················· | ···················· | ···················· |
| X | 991 | hCoV-19/England/204991191/2020\|EPI_ISL_723033\|2020 | ···················· | ···················· | ···················· | ···················· | ···················· | ···················· |
| X | 992 | hCoV-19/England/204991196/2020\|EPI_ISL_723034\|2020 | ···················· | ···················· | ···················· | ···················· | ···················· | ···················· |
| X | 993 | hCoV-19/England/204991204/2020\|EPI_ISL_723035\|2020 | ···················· | ···················· | ···················· | ···················· | ···················· | ···················· |
| X | 994 | hCoV-19/England/205020023/2020\|EPI_ISL_723036\|2020 | ···················· | ···················· | ···················· | ···················· | ···················· | ···················· |
| X | 995 | hCoV-19/England/205020035/2020\|EPI_ISL_723037\|2020 | ···················· | ···················· | ···················· | ···················· | ···················· | ···················· |
| X | 996 | hCoV-19/England/205020335/2020\|EPI_ISL_723038\|2020 | ···················· | ···················· | ···················· | ···················· | ···················· | ···················· |
| X | 997 | hCoV-19/England/205020748/2020\|EPI_ISL_723039\|2020 | ···················· | ···················· | ···················· | ···················· | ···················· | ···················· |
| X | 998 | hCoV-19/England/205020752/2020\|EPI_ISL_723040\|2020 | ···················· | ···················· | ···················· | ···················· | ···················· | ···················· |
| X | 999 | hCoV-19/England/205020756/2020\|EPI_ISL_723041\|2020 | ···················· | ···················· | ···················· | ···················· | ···················· | ···················· |
| X | 1000 | hCoV-19/England/205060152/2020\|EPI_ISL_723043\|2020 | ···················· | ···················· | ···················· | ···················· | ···················· | ···················· |
| X | 1001 | hCoV-19/England/EXET-13E527/2020\|EPI_ISL_724326\|20 | ···················· | ···················· | ···················· | ···················· | ···················· | ···················· |
| X | 1002 | hCoV-19/England/EXET-13E624/2020\|EPI_ISL_724337\|20 | ···················· | ···················· | ···················· | ···················· | ···················· | ···················· |
| X | 1003 | hCoV-19/England/EXET-13E651/2020\|EPI_ISL_724339\|20 | ···················· | ·A·················· | ···················· | ···················· | ······T············· | ···················· |
| X | 1004 | hCoV-19/England/EXET-13E660/2020\|EPI_ISL_724340\|20 | ···················· | ···················· | ···················· | ···················· | ···················· | ···················· |
| X | 1005 | hCoV-19/England/EXET-13E67F/2020\|EPI_ISL_724341\|20 | ···················· | ···················· | ···················· | ···················· | ······T············· | ···················· |
| X | 1006 | hCoV-19/England/LIVE-DCA3C0/2020\|EPI_ISL_724356\|20 | ···················· | ···················· | ···················· | ···················· | ···················· | ···················· |
| X | 1007 | hCoV-19/England/LIVE-DCA3EE/2020\|EPI_ISL_724357\|20 | ···················· | ···················· | ···················· | ···················· | ···················· | ···················· |
| X | 1008 | hCoV-19/England/LIVE-DCA436/2020\|EPI_ISL_724361\|20 | ···················· | ···················· | ···················· | ···················· | ···················· | ···················· |
| X | 1009 | hCoV-19/England/LIVE-DCA472/2020\|EPI_ISL_724364\|20 | ···················· | ···················· | ···················· | ···················· | ···················· | ···················· |
| X | 1010 | hCoV-19/Wales/LIVE-DCA4FA/2020\|EPI_ISL_724365\|2020 | ···················· | ···················· | ···················· | ···················· | ···················· | ···················· |
| X | 1011 | hCoV-19/England/LIVE-DCA506/2020\|EPI_ISL_724366\|20 | ···················· | ···················· | ···················· | ···················· | ···················· | ···················· |
| X | 1012 | hCoV-19/England/LIVE-DCA5AC/2020\|EPI_ISL_724367\|20 | ···················· | ···················· | ···················· | ···················· | ···················· | ···················· |
| X | 1013 | hCoV-19/England/LIVE-DCA5D9/2020\|EPI_ISL_724368\|20 | ···················· | ···················· | ···················· | ···················· | ···················· | ···················· |
| X | 1014 | hCoV-19/England/LIVE-DCA5F7/2020\|EPI_ISL_724369\|20 | ···················· | ···················· | ···················· | ···················· | ···················· | ···················· |
| X | 1015 | hCoV-19/England/LIVE-DCA603/2020\|EPI_ISL_724370\|20 | ···················· | ···················· | ···················· | ···················· | ···················· | ···················· |
| X | 1016 | hCoV-19/England/LIVE-DCA67C/2020\|EPI_ISL_724372\|20 | ···················· | ···················· | ···················· | ···················· | ···················· | ···················· |
| X | 1017 | hCoV-19/England/LIVE-DCA6F4/2020\|EPI_ISL_724378\|20 | ···················· | ···················· | ···················· | ···················· | ···················· | ···················· |
| X | 1018 | hCoV-19/England/LIVE-DCA74C/2020\|EPI_ISL_724380\|20 | ···················· | ···················· | ···················· | ···················· | ···················· | ···················· |
| X | 1019 | hCoV-19/England/LIVE-DCA76A/2020\|EPI_ISL_724382\|20 | ···················· | ···················· | ···················· | ···················· | ···················· | ···················· |
| X | 1020 | hCoV-19/England/LIVE-DCA779/2020\|EPI_ISL_724383\|20 | ···················· | ···················· | ···················· | ···················· | ···················· | ···················· |
| X | 1021 | hCoV-19/England/LIVE-DCA788/2020\|EPI_ISL_724384\|20 | ···················· | ···················· | ···················· | ···················· | ···A················ | ···················· |
| X | 1022 | hCoV-19/England/LIVE-DCA7F1/2020\|EPI_ISL_724387\|20 | ···················· | ···················· | ···················· | ···················· | ······T············· | ···················· |
| X | 1023 | hCoV-19/England/LIVE-DCA876/2020\|EPI_ISL_724388\|20 | ···················· | ···················· | ···················· | ···················· | ···················· | ···················· |
| X | 1024 | hCoV-19/England/LIVE-DCA894/2020\|EPI_ISL_724389\|20 | ···················· | ···················· | ···················· | ···················· | ···················· | ···················· |
| X | 1025 | hCoV-19/Denmark/DCGC-13710/2020\|EPI_ISL_712400\|202 | ···················· | ···················· | ···················· | ···················· | ···················· | ···················· |
| X | 1026 | hCoV-19/Denmark/DCGC-13722/2020\|EPI_ISL_712401\|202 | ···················· | ···················· | ···················· | ···················· | ···················· | ···················· |
| X | 1027 | hCoV-19/Denmark/DCGC-13724/2020\|EPI_ISL_712402\|202 | ···················· | ···················· | ···················· | ···················· | ···················· | ···················· |
| X | 1028 | hCoV-19/Denmark/DCGC-13726/2020\|EPI_ISL_712403\|202 | ···················· | ···················· | ···················· | ···················· | ···················· | ···················· |
| X | 1029 | hCoV-19/Denmark/DCGC-13742/2020\|EPI_ISL_712404\|202 | ···················· | ···················· | ···················· | ···················· | ···················· | ···················· |
| X | 1030 | hCoV-19/Denmark/DCGC-13759/2020\|EPI_ISL_712405\|202 | ···················· | ···················· | ···················· | ···················· | ···C················ | ···················· |
| X | 1031 | hCoV-19/Denmark/DCGC-13770/2020\|EPI_ISL_712406\|202 | ···················· | ···················· | ···················· | ···················· | ···················· | ···················· |
| X | 1032 | hCoV-19/Denmark/DCGC-13773/2020\|EPI_ISL_712407\|202 | ···················· | ···················· | ···················· | ···················· | ···················· | ···················· |
| X | 1033 | hCoV-19/Denmark/DCGC-13775/2020\|EPI_ISL_712408\|202 | ···················· | ···················· | ···················· | ···················· | ···················· | ···················· |
| X | 1034 | hCoV-19/Denmark/DCGC-13809/2020\|EPI_ISL_712409\|202 | ···················· | ···················· | ···················· | ···················· | ···················· | ···················· |
| X | 1035 | hCoV-19/Denmark/DCGC-13863/2020\|EPI_ISL_712415\|202 | ···················· | ···················· | ···················· | ···················· | ···················· | ···················· |
| X | 1036 | hCoV-19/Denmark/DCGC-15217/2020\|EPI_ISL_713700\|202 | ···················· | ···················· | ···················· | ···················· | ···················· | ···················· |
| X | 1037 | hCoV-19/Denmark/DCGC-15218/2020\|EPI_ISL_713701\|202 | ···················· | ···················· | ···················· | ···················· | ···················· | ···················· |
| X | 1038 | hCoV-19/Denmark/DCGC-15219/2020\|EPI_ISL_713702\|202 | ···················· | ···················· | ···················· | ···················· | ···················· | ···················· |
| X | 1039 | hCoV-19/Denmark/DCGC-15220/2020\|EPI_ISL_713703\|202 | ···················· | ·A·················· | ···················· | ···················· | ···················· | ···A················ |
| X | 1040 | hCoV-19/Denmark/DCGC-15221/2020\|EPI_ISL_713704\|202 | ···················· | ···················· | ···················· | ···················· | ···················· | ···················· |
| X | 1041 | hCoV-19/Denmark/DCGC-15222/2020\|EPI_ISL_713705\|202 | ···················· | ···················· | ···················· | ···················· | ···················· | ···················· |
| X | 1042 | hCoV-19/Denmark/DCGC-15224/2020\|EPI_ISL_713706\|202 | ···················· | ···················· | ···················· | ···················· | ···················· | ···················· |
| X | 1043 | hCoV-19/Denmark/DCGC-15225/2020\|EPI_ISL_713707\|202 | ···················· | ···················· | ···················· | ···················· | ···················· | ···················· |
| X | 1044 | hCoV-19/Denmark/DCGC-15227/2020\|EPI_ISL_713708\|202 | ···················· | ···················· | ···················· | ···················· | ···················· | ···················· |
| X | 1045 | hCoV-19/Denmark/DCGC-15228/2020\|EPI_ISL_713709\|202 | ···················· | ···················· | ···················· | ···················· | ···················· | ···················· |
| X | 1046 | hCoV-19/Denmark/DCGC-15230/2020\|EPI_ISL_713710\|202 | ···················· | ···················· | ···················· | ···················· | ···················· | ···················· |

| | | | | | | | | |
|---|---|---|---|---|---|---|---|---|
| X | 1047 | hCoV-19/Denmark/DCGC-15232/2020\|EPI_ISL_713711\|202 | ···················· | ···················· | ····················· | ···················· | ···················· | ···················· |
| X | 1048 | hCoV-19/Denmark/DCGC-15233/2020\|EPI_ISL_713712\|202 | ···················· | ···················· | ····················· | ···················· | ···················· | ···················· |
| X | 1049 | hCoV-19/Denmark/DCGC-15234/2020\|EPI_ISL_713713\|202 | ···················· | ···················· | ····················· | ···················· | ···················· | ···················· |
| X | 1050 | hCoV-19/Denmark/DCGC-15235/2020\|EPI_ISL_713714\|202 | ···················· | ···················· | ····················· | ···················· | ···················· | ···················· |
| X | 1051 | hCoV-19/Denmark/DCGC-15239/2020\|EPI_ISL_713715\|202 | ···················· | ···················· | ····················· | ···················· | ···················· | ···················· |
| X | 1052 | hCoV-19/Denmark/DCGC-15240/2020\|EPI_ISL_713716\|202 | ···················· | ···················· | ····················· | ···················· | ···················· | ···················· |
| X | 1053 | hCoV-19/Denmark/DCGC-15247/2020\|EPI_ISL_713717\|202 | ···················· | ···················· | ····················· | ···················· | ···················· | ···················· |
| X | 1054 | hCoV-19/Denmark/DCGC-15248/2020\|EPI_ISL_713718\|202 | ···················· | ···················· | ····················· | ···················· | ···················· | ···················· |
| X | 1055 | hCoV-19/Denmark/DCGC-15249/2020\|EPI_ISL_713719\|202 | ···················· | ···················· | ····················· | ···················· | ···················· | ···················· |
| X | 1056 | hCoV-19/Denmark/DCGC-15251/2020\|EPI_ISL_713720\|202 | ···················· | ···················· | ····················· | ···················· | ···················· | ···················· |
| X | 1057 | hCoV-19/Denmark/DCGC-15253/2020\|EPI_ISL_713721\|202 | ···················· | ·············A······ | ····················· | ···················· | ···················· | ···················· |
| X | 1058 | hCoV-19/Denmark/DCGC-15254/2020\|EPI_ISL_713722\|202 | ···················· | ·A·················· | ····················· | ···················· | ···················· | ···A················ |
| X | 1059 | hCoV-19/Denmark/DCGC-15255/2020\|EPI_ISL_713723\|202 | ···················· | ···················· | ····················· | ···················· | ···················· | ···················· |
| X | 1060 | hCoV-19/Denmark/DCGC-15256/2020\|EPI_ISL_713724\|202 | ···················· | ···················· | ····················· | ···················· | ···················· | ···················· |
| X | 1061 | hCoV-19/Denmark/DCGC-15257/2020\|EPI_ISL_713725\|202 | ···················· | ···················· | ····················· | ···················· | ···················· | ···················· |
| X | 1062 | hCoV-19/Denmark/DCGC-15258/2020\|EPI_ISL_713726\|202 | ···················· | ···················· | ····················· | ···················· | ···················· | ···················· |
| X | 1063 | hCoV-19/Denmark/DCGC-15259/2020\|EPI_ISL_713727\|202 | ···················· | ···················· | ····················· | ···················· | ···················· | ···················· |
| X | 1064 | hCoV-19/Denmark/DCGC-15260/2020\|EPI_ISL_713728\|202 | ···················· | ···················· | ····················· | ···················· | ···················· | ···················· |
| X | 1065 | hCoV-19/Denmark/DCGC-15261/2020\|EPI_ISL_713729\|202 | ···················· | ···················· | ····················· | ···················· | ···················· | ···················· |
| X | 1066 | hCoV-19/Denmark/DCGC-15262/2020\|EPI_ISL_713730\|202 | ···················· | ···················· | ····················· | ···················· | ···················· | ···················· |
| X | 1067 | hCoV-19/Denmark/DCGC-15266/2020\|EPI_ISL_713731\|202 | ···················· | ···················· | ····················· | ···················· | ···················· | ···················· |
| X | 1068 | hCoV-19/Denmark/DCGC-15267/2020\|EPI_ISL_713732\|202 | ···················· | ···················· | ····················· | ···················· | ···················· | ···················· |
| X | 1069 | hCoV-19/Denmark/DCGC-15269/2020\|EPI_ISL_713733\|202 | ···················· | ···················· | ····················· | ···················· | ···················· | ···················· |
| X | 1070 | hCoV-19/Denmark/DCGC-15270/2020\|EPI_ISL_713734\|202 | ···················· | ···················· | ····················· | ···················· | ···················· | ···················· |
| X | 1071 | hCoV-19/Denmark/DCGC-15271/2020\|EPI_ISL_713735\|202 | ···················· | ···················· | ····················· | ···················· | ···················· | ···················· |
| X | 1072 | hCoV-19/Denmark/DCGC-15272/2020\|EPI_ISL_713736\|202 | ···················· | ···················· | ····················· | ···················· | ···················· | ···················· |
| X | 1073 | hCoV-19/Denmark/DCGC-15274/2020\|EPI_ISL_713737\|202 | ···················· | ···················· | ····················· | ···················· | ···················· | ···················· |
| X | 1074 | hCoV-19/Denmark/DCGC-15276/2020\|EPI_ISL_713738\|202 | ···················· | ···················· | ····················· | ···················· | ···················· | ···················· |
| X | 1075 | hCoV-19/Denmark/DCGC-15279/2020\|EPI_ISL_713739\|202 | ···················· | ···················· | ····················· | ···················· | ···················· | ···················· |
| X | 1076 | hCoV-19/Denmark/DCGC-15291/2020\|EPI_ISL_713746\|202 | ···················· | ···················· | ····················· | ···················· | ···················· | ···················· |
| X | 1077 | hCoV-19/Denmark/DCGC-15294/2020\|EPI_ISL_713747\|202 | ···················· | ···················· | ····················· | ···················· | ···················· | ···················· |
| X | 1078 | hCoV-19/Denmark/DCGC-15313/2020\|EPI_ISL_713759\|202 | ···················· | ···················· | ····················· | ···················· | ···················· | ···················· |
| X | 1079 | hCoV-19/Denmark/DCGC-15314/2020\|EPI_ISL_713760\|202 | ···················· | ···················· | ····················· | ···················· | ···················· | ···················· |
| X | 1080 | hCoV-19/Denmark/DCGC-15317/2020\|EPI_ISL_713761\|202 | ···················· | ···················· | ····················· | ···················· | ···················· | ···················· |
| X | 1081 | hCoV-19/Denmark/DCGC-15320/2020\|EPI_ISL_713762\|202 | ···················· | ···················· | ····················· | ···················· | ···················· | ···················· |
| X | 1082 | hCoV-19/Denmark/DCGC-15321/2020\|EPI_ISL_713763\|202 | ···················· | ···················· | ····················· | ···················· | ···················· | ···················· |
| X | 1083 | hCoV-19/Denmark/DCGC-15322/2020\|EPI_ISL_713764\|202 | ···················· | ···················· | ····················· | ···················· | ···················· | ···················· |
| X | 1084 | hCoV-19/Denmark/DCGC-15324/2020\|EPI_ISL_713765\|202 | ···················· | ···················· | ····················· | ···················· | ···················· | ···················· |
| X | 1085 | hCoV-19/Denmark/DCGC-15330/2020\|EPI_ISL_713766\|202 | ···················· | ···················· | ····················· | ···················· | ···················· | ···················· |
| X | 1086 | hCoV-19/Denmark/DCGC-15331/2020\|EPI_ISL_713767\|202 | ···················· | ···················· | ····················· | ···················· | ···················· | ···················· |
| X | 1087 | hCoV-19/Denmark/DCGC-15333/2020\|EPI_ISL_713768\|202 | ···················· | ···················· | ····················· | ···················· | ···················· | ···················· |
| X | 1088 | hCoV-19/Denmark/DCGC-15337/2020\|EPI_ISL_713769\|202 | ···················· | ···················· | ····················· | ···················· | ···················· | ···················· |
| X | 1089 | hCoV-19/Denmark/DCGC-15338/2020\|EPI_ISL_713770\|202 | ···················· | ···················· | ····················· | ···················· | ···················· | ···················· |
| X | 1090 | hCoV-19/Denmark/DCGC-15339/2020\|EPI_ISL_713771\|202 | ···················· | ···················· | ····················· | ···················· | ···················· | ···················· |
| X | 1091 | hCoV-19/Denmark/DCGC-15340/2020\|EPI_ISL_713772\|202 | ···················· | ···················· | ····················· | ···················· | ···················· | ···················· |
| X | 1092 | hCoV-19/Denmark/DCGC-15341/2020\|EPI_ISL_713773\|202 | ···················· | ···················· | ····················· | ···················· | ···················· | ···················· |
| X | 1093 | hCoV-19/Denmark/DCGC-15342/2020\|EPI_ISL_713774\|202 | ···················· | ···················· | ····················· | ···················· | ···················· | ···················· |
| X | 1094 | hCoV-19/Denmark/DCGC-15343/2020\|EPI_ISL_713775\|202 | ···················· | ···················· | ····················· | ···················· | ···················· | ···················· |
| X | 1095 | hCoV-19/Denmark/DCGC-15348/2020\|EPI_ISL_713776\|202 | ···················· | ··········A········· | ····················· | ···················· | ···················· | ···················· |
| X | 1096 | hCoV-19/Denmark/DCGC-15349/2020\|EPI_ISL_713777\|202 | ···················· | ···················· | ····················· | ···················· | ···················· | ···················· |
| X | 1097 | hCoV-19/Denmark/DCGC-15351/2020\|EPI_ISL_713778\|202 | ···················· | ···················· | ····················· | ···················· | ···················· | ···················· |
| X | 1098 | hCoV-19/Denmark/DCGC-15352/2020\|EPI_ISL_713779\|202 | ···················· | ···················· | ····················· | ···················· | ···················· | ···················· |
| X | 1099 | hCoV-19/England/CAMC-C46837/2020\|EPI_ISL_718504\|20 | ········T··········· | ···················· | ····················· | ···················· | ···················· | ···················· |
| X | 1100 | hCoV-19/England/CAMC-C44D76/2020\|EPI_ISL_718506\|20 | ···················· | ···················· | ····················· | ···················· | ···················· | ···················· |
| X | 1101 | hCoV-19/England/CAMC-C46A31/2020\|EPI_ISL_718509\|20 | ···················· | ···················· | ····················· | ···················· | ···················· | ···················· |
| X | 1102 | hCoV-19/England/CAMC-C46AE6/2020\|EPI_ISL_718517\|20 | ···················· | ···················· | ····················· | ···················· | ···················· | ···················· |
| X | 1103 | hCoV-19/England/CAMC-C44D67/2020\|EPI_ISL_718525\|20 | ···················· | ···················· | ····················· | ···················· | ···················· | ···················· |
| X | 1104 | hCoV-19/England/CAMC-C45FBB/2020\|EPI_ISL_718526\|20 | ···················· | ···················· | ····················· | ···················· | ···················· | ···················· |
| X | 1105 | hCoV-19/England/CAMC-C44B7C/2020\|EPI_ISL_718527\|20 | ···················· | ···················· | ····················· | ···················· | ···················· | ···················· |
| X | 1106 | hCoV-19/England/CAMC-C3FD6F/2020\|EPI_ISL_718529\|20 | ···················· | ···················· | ····················· | ···················· | ···················· | ···················· |
| X | 1107 | hCoV-19/England/CAMC-C3F6DD/2020\|EPI_ISL_718536\|20 | ···················· | ·A·················· | ····················· | ···················· | ···················· | ···A················ |
| X | 1108 | hCoV-19/England/CAMC-C3FD32/2020\|EPI_ISL_718542\|20 | ···················· | ···················· | ····················· | ···················· | ···················· | ···················· |
| X | 1109 | hCoV-19/England/CAMC-C4663D/2020\|EPI_ISL_718543\|20 | ···················· | ···················· | ····················· | ···················· | ···················· | ···················· |
| X | 1110 | hCoV-19/England/CAMC-C3F6A0/2020\|EPI_ISL_718544\|20 | ···················· | ···················· | ····················· | ···················· | ···················· | ···················· |
| X | 1111 | hCoV-19/England/CAMC-C46AD7/2020\|EPI_ISL_718547\|20 | ···················· | ···················· | ····················· | ···················· | ···················· | ···················· |
| X | 1112 | hCoV-19/England/CAMC-C44BC7/2020\|EPI_ISL_718553\|20 | ···················· | ···················· | ····················· | ···················· | ···················· | ···················· |
| X | 1113 | hCoV-19/England/CAMC-C46AAA/2020\|EPI_ISL_718556\|20 | ···················· | ···················· | ····················· | ···················· | ···················· | ···················· |
| X | 1114 | hCoV-19/England/CAMC-C46864/2020\|EPI_ISL_718558\|20 | ···················· | ···················· | ····················· | ···················· | ···················· | ···················· |
| X | 1115 | hCoV-19/England/CAMC-C402C6/2020\|EPI_ISL_718561\|20 | ···················· | ·A·················· | ····················· | ···················· | ···················· | ···A················ |
| X | 1116 | hCoV-19/England/CAMC-C45F8E/2020\|EPI_ISL_718579\|20 | ···················· | ···················· | ····················· | ···················· | ···················· | ···················· |
| X | 1117 | hCoV-19/England/CAMC-C479F7/2020\|EPI_ISL_718580\|20 | ···················· | ···················· | ····················· | ···················· | ···················· | ···················· |
| X | 1118 | hCoV-19/England/CAMC-C452B2/2020\|EPI_ISL_718583\|20 | ···················· | ···················· | ····················· | ···················· | ···················· | ···················· |
| X | 1119 | hCoV-19/England/CAMC-C45EBE/2020\|EPI_ISL_718586\|20 | ···················· | ···················· | ····················· | ···················· | ······T············· | ···················· |
| X | 1120 | hCoV-19/England/CAMC-C45FD9/2020\|EPI_ISL_718587\|20 | ···················· | ···················· | ····················· | ···················· | ···················· | ···················· |
| X | 1121 | hCoV-19/England/CAMC-C403A5/2020\|EPI_ISL_718592\|20 | ···················· | ···················· | ····················· | ···················· | ···················· | ···················· |
| X | 1122 | hCoV-19/England/CAMC-C45C3C/2020\|EPI_ISL_718594\|20 | ···················· | ···················· | ····················· | ···················· | ···················· | ···················· |
| X | 1123 | hCoV-19/England/CAMC-C40396/2020\|EPI_ISL_718595\|20 | ···················· | ···················· | ····················· | ···················· | ···················· | ···················· |
| X | 1124 | hCoV-19/England/QEUH-BA0DB5/2020\|EPI_ISL_719800\|20 | ···················· | ···················· | ····················· | ···················· | ···················· | ···················· |
| X | 1125 | hCoV-19/England/QEUH-BA0DA6/2020\|EPI_ISL_719802\|20 | ···················· | ·A·················· | ····················· | ···················· | ···················· | ···················· |
| X | 1126 | hCoV-19/England/QEUH-BA05CC/2020\|EPI_ISL_719803\|20 | ···················· | ···················· | ····················· | ···················· | ···················· | ···················· |

| | | | | | | | | |
|---|---|---|---|---|---|---|---|---|
| X | 1127 | hCoV-19/England/QEUH-B9FFFF/2020\|EPI_ISL_719804\|20 | ···················· | ···················· | ···················· | ···················· | ···················· | ···················· |
| X | 1128 | hCoV-19/England/QEUH-BA1798/2020\|EPI_ISL_719805\|20 | ···················· | ···················· | ···················· | ···················· | ···················· | ···················· |
| X | 1129 | hCoV-19/England/QEUH-BA0483/2020\|EPI_ISL_719806\|20 | ···················· | ···················· | ···················· | ···················· | ···················· | ···················· |
| X | 1130 | hCoV-19/England/QEUH-BA0043/2020\|EPI_ISL_719807\|20 | ···················· | ···················· | ···················· | ···················· | ···················· | ···················· |
| X | 1131 | hCoV-19/England/QEUH-BA1F72/2020\|EPI_ISL_719808\|20 | ···················· | ···················· | ···················· | ···················· | ···················· | ···················· |
| X | 1132 | hCoV-19/England/QEUH-BA2393/2020\|EPI_ISL_719809\|20 | ···················· | ···················· | ···················· | ···················· | ···················· | ···················· |
| X | 1133 | hCoV-19/England/QEUH-BA2120/2020\|EPI_ISL_719810\|20 | ···················· | ···················· | ···················· | ···················· | ···················· | ···················· |
| X | 1134 | hCoV-19/England/QEUH-BA183B/2020\|EPI_ISL_719811\|20 | ···················· | ···················· | ···················· | ···················· | ···················· | ···················· |
| X | 1135 | hCoV-19/England/QEUH-B9F135/2020\|EPI_ISL_719812\|20 | ···················· | ···················· | ···················· | ···················· | ···················· | ···················· |
| X | 1136 | hCoV-19/England/QEUH-BA1BF6/2020\|EPI_ISL_719813\|20 | ···················· | ···················· | ···················· | ···················· | ···················· | ···················· |
| X | 1137 | hCoV-19/England/QEUH-B9EA2C/2020\|EPI_ISL_719814\|20 | ···················· | ···················· | ···················· | ···················· | ···················· | ···················· |
| X | 1138 | hCoV-19/England/QEUH-B9E734/2020\|EPI_ISL_719815\|20 | ···················· | ···················· | ···················· | ···················· | ···················· | ···················· |
| X | 1139 | hCoV-19/England/QEUH-BA230C/2020\|EPI_ISL_719817\|20 | ···················· | ···················· | ···················· | ···················· | ···················· | ···················· |
| X | 1140 | hCoV-19/England/QEUH-B9EB65/2020\|EPI_ISL_719818\|20 | ···················· | ···················· | ···················· | ···················· | ···················· | ···················· |
| X | 1141 | hCoV-19/England/QEUH-BA1C3F/2020\|EPI_ISL_719819\|20 | ···················· | ···················· | ···················· | ···················· | ···················· | ···A················ |
| X | 1142 | hCoV-19/England/QEUH-B9E7CB/2020\|EPI_ISL_719821\|20 | ···················· | ···················· | ···················· | ···················· | ···················· | ···················· |
| X | 1143 | hCoV-19/England/QEUH-BA1FFA/2020\|EPI_ISL_719822\|20 | ···················· | ···················· | ···················· | ···················· | ···················· | ···················· |
| X | 1144 | hCoV-19/England/QEUH-BA1C02/2020\|EPI_ISL_719823\|20 | ···················· | ···················· | ···················· | ···················· | ···················· | ···················· |
| X | 1145 | hCoV-19/England/QEUH-B9F117/2020\|EPI_ISL_719829\|20 | ···················· | ···················· | ···················· | ···················· | ···················· | ···················· |
| X | 1146 | hCoV-19/England/QEUH-B9EAA4/2020\|EPI_ISL_719834\|20 | ···················· | ···················· | ···················· | ···················· | ···················· | ···················· |
| X | 1147 | hCoV-19/England/QEUH-BA1FBE/2020\|EPI_ISL_719839\|20 | ···················· | ···················· | ···················· | ···················· | ···················· | ···················· |
| X | 1148 | hCoV-19/England/QEUH-B9ED6F/2020\|EPI_ISL_719841\|20 | ···················· | ···················· | ···················· | ···················· | ···················· | ···················· |
| X | 1149 | hCoV-19/England/QEUH-B9EB29/2020\|EPI_ISL_719846\|20 | ···················· | ···················· | ···················· | ···················· | ···················· | ···················· |
| X | 1150 | hCoV-19/England/QEUH-BA17C5/2020\|EPI_ISL_719847\|20 | ···················· | ···················· | ···················· | ···················· | ···················· | ···················· |
| X | 1151 | hCoV-19/England/QEUH-B9F047/2020\|EPI_ISL_719848\|20 | ···················· | ···················· | ···················· | ···················· | ···················· | ···A················ |
| X | 1152 | hCoV-19/England/QEUH-B9ECF9/2020\|EPI_ISL_719849\|20 | ···················· | ···················· | ···················· | ···················· | ···················· | ···················· |
| X | 1153 | hCoV-19/England/QEUH-BA18C2/2020\|EPI_ISL_719850\|20 | ···················· | ···················· | ···················· | ···················· | ···················· | ···················· |
| X | 1154 | hCoV-19/England/QEUH-B9F1CC/2020\|EPI_ISL_719852\|20 | ···················· | ···················· | ···················· | ···················· | ···················· | ···················· |
| X | 1155 | hCoV-19/England/QEUH-BA2014/2020\|EPI_ISL_719856\|20 | ···················· | ···················· | ···················· | ···················· | ···················· | ···················· |
| X | 1156 | hCoV-19/England/QEUH-BA1B7E/2020\|EPI_ISL_719857\|20 | ···················· | ···················· | ···················· | ···················· | ···················· | ···················· |
| X | 1157 | hCoV-19/England/QEUH-B9ED7E/2020\|EPI_ISL_719858\|20 | ···················· | ···················· | ···················· | ···················· | ···················· | ···················· |
| X | 1158 | hCoV-19/England/QEUH-B9F083/2020\|EPI_ISL_719861\|20 | ···················· | ···················· | ···················· | ···················· | ···················· | ···················· |
| X | 1159 | hCoV-19/England/QEUH-BA1C99/2020\|EPI_ISL_719862\|20 | ···················· | ···················· | ···················· | ···················· | ···················· | ···················· |
| X | 1160 | hCoV-19/England/QEUH-BA20AB/2020\|EPI_ISL_719864\|20 | ···················· | ···················· | ···················· | ···················· | ···················· | ···················· |
| X | 1161 | hCoV-19/England/QEUH-BA231B/2020\|EPI_ISL_719865\|20 | ···················· | ···················· | ···················· | ···················· | ···················· | ···················· |
| X | 1162 | hCoV-19/England/QEUH-B9F153/2020\|EPI_ISL_719866\|20 | ···················· | ···················· | ···················· | ···················· | ···················· | ···················· |
| X | 1163 | hCoV-19/England/QEUH-BA18D1/2020\|EPI_ISL_719867\|20 | ···················· | ···················· | ···················· | ···················· | ···················· | ···················· |
| X | 1164 | hCoV-19/England/QEUH-B9E840/2020\|EPI_ISL_719868\|20 | ···················· | ···················· | ···················· | ···················· | ···················· | ···················· |
| X | 1165 | hCoV-19/England/QEUH-BA271F/2020\|EPI_ISL_719870\|20 | ···················· | ···················· | ···················· | ···················· | ······T············· | ···················· |
| X | 1166 | hCoV-19/England/QEUH-B9E6FB/2020\|EPI_ISL_719871\|20 | ···················· | ···················· | ···················· | ···················· | ···················· | ···················· |
| X | 1167 | hCoV-19/England/QEUH-BA225A/2020\|EPI_ISL_719872\|20 | ···················· | ·A·················· | ···················· | ···················· | ···················· | ···A················ |
| X | 1168 | hCoV-19/England/QEUH-B9E86E/2020\|EPI_ISL_719873\|20 | ···················· | ···················· | ···················· | ···················· | ···················· | ···················· |
| X | 1169 | hCoV-19/England/QEUH-B9EDE7/2020\|EPI_ISL_719875\|20 | ···················· | ···················· | ···················· | ···················· | ···················· | ···················· |
| X | 1170 | hCoV-19/England/QEUH-BA218A/2020\|EPI_ISL_719876\|20 | ···················· | ···················· | ···················· | ···················· | ···················· | ···················· |
| X | 1171 | hCoV-19/England/QEUH-B9EE7B/2020\|EPI_ISL_719877\|20 | ···················· | ···················· | ···················· | ···················· | ···················· | ···················· |
| X | 1172 | hCoV-19/England/QEUH-BA27B5/2020\|EPI_ISL_719878\|20 | ···················· | ···················· | ···················· | ···················· | ···················· | ···················· |
| X | 1173 | hCoV-19/England/QEUH-BA2296/2020\|EPI_ISL_719880\|20 | ···················· | ·A·················· | ···················· | ···················· | ···················· | ···A················ |
| X | 1174 | hCoV-19/England/QEUH-B9F0ED/2020\|EPI_ISL_719881\|20 | ···················· | ···················· | ···················· | ···················· | ···················· | ···················· |
| X | 1175 | hCoV-19/England/QEUH-B9E761/2020\|EPI_ISL_719883\|20 | ···················· | ···················· | ···················· | ···················· | ···················· | ···················· |
| X | 1176 | hCoV-19/England/QEUH-B9E7BC/2020\|EPI_ISL_719884\|20 | ···················· | ···················· | ···················· | ···················· | ···················· | ···················· |
| X | 1177 | hCoV-19/England/QEUH-B9EE20/2020\|EPI_ISL_719885\|20 | ···················· | ···················· | ···················· | ···················· | ···················· | ···················· |
| X | 1178 | hCoV-19/Wales/QEUH-BA17F2/2020\|EPI_ISL_719887\|2020 | ···················· | ···················· | ···················· | ···················· | ···················· | ···················· |
| X | 1179 | hCoV-19/England/QEUH-BA22C3/2020\|EPI_ISL_719889\|20 | ···················· | ···················· | ···················· | ···················· | ···················· | ···················· |
| X | 1180 | hCoV-19/England/QEUH-BA190B/2020\|EPI_ISL_719890\|20 | ···················· | ···················· | ···················· | ···················· | ···················· | ···················· |
| X | 1181 | hCoV-19/England/QEUH-B9E8C8/2020\|EPI_ISL_719891\|20 | ···················· | ···················· | ···················· | ···················· | ···················· | ···················· |
| X | 1182 | hCoV-19/England/QEUH-BA1868/2020\|EPI_ISL_719892\|20 | ···················· | ···················· | ···················· | ···················· | ···················· | ···················· |
| X | 1183 | hCoV-19/England/QEUH-BA27D3/2020\|EPI_ISL_719894\|20 | ···················· | ···················· | ···················· | ···················· | ···················· | ···················· |
| X | 1184 | hCoV-19/England/QEUH-B9E7F8/2020\|EPI_ISL_719896\|20 | ···················· | ···················· | ···················· | ···················· | ···················· | ···················· |
| X | 1185 | hCoV-19/England/QEUH-B9EA95/2020\|EPI_ISL_719898\|20 | ···················· | ···················· | ···················· | ···················· | ···················· | ···················· |
| X | 1186 | hCoV-19/England/MILK-C0067B/2020\|EPI_ISL_720804\|20 | ···················· | ···················· | ···················· | ···················· | ···················· | ···················· |
| X | 1187 | hCoV-19/England/QEUH-C01364/2020\|EPI_ISL_720805\|20 | ···················· | ···················· | ···················· | ···················· | ···················· | ···················· |
| X | 1188 | hCoV-19/England/MILK-BFD566/2020\|EPI_ISL_720808\|20 | ···················· | ···················· | ···················· | ···················· | ···················· | ···················· |
| X | 1189 | hCoV-19/England/MILK-BFCD7E/2020\|EPI_ISL_720809\|20 | ···················· | ···················· | ···················· | ···················· | ···················· | ···················· |
| X | 1190 | hCoV-19/England/ALDP-C04A99/2020\|EPI_ISL_720810\|20 | ···················· | ···················· | ···················· | ···················· | ···················· | ···················· |
| X | 1191 | hCoV-19/England/QEUH-C013BF/2020\|EPI_ISL_720811\|20 | ···················· | ···················· | ···················· | ···················· | ···················· | ···················· |
| X | 1192 | hCoV-19/England/MILK-BFCCDB/2020\|EPI_ISL_720812\|20 | ···················· | ···················· | ···················· | ···················· | ···················· | ···················· |
| X | 1193 | hCoV-19/England/ALDP-C049E7/2020\|EPI_ISL_720816\|20 | ···················· | ···················· | ···················· | ···················· | ···················· | ···················· |
| X | 1194 | hCoV-19/England/MILK-BFF4C1/2020\|EPI_ISL_720817\|20 | ···················· | ···················· | ···················· | ···················· | ···················· | ···················· |
| X | 1195 | hCoV-19/England/QEUH-C013FB/2020\|EPI_ISL_720819\|20 | ···················· | ···················· | ···················· | ···················· | ···················· | ···················· |
| X | 1196 | hCoV-19/England/MILK-C008B1/2020\|EPI_ISL_720820\|20 | ···················· | ···················· | ···················· | ···················· | ···················· | ···················· |
| X | 1197 | hCoV-19/England/MILK-BFF555/2020\|EPI_ISL_720822\|20 | ···················· | ···················· | ···················· | ···················· | ···················· | ···················· |
| X | 1198 | hCoV-19/England/ALDP-C04AC6/2020\|EPI_ISL_720823\|20 | ···················· | ·A·················· | ···················· | ···················· | ···················· | ···················· |
| X | 1199 | hCoV-19/England/MILK-BFF652/2020\|EPI_ISL_720824\|20 | ···················· | ···················· | ···················· | ···················· | ···················· | ···················· |
| X | 1200 | hCoV-19/England/ALDP-C04B1E/2020\|EPI_ISL_720827\|20 | ···················· | ···················· | ···················· | ···················· | ···················· | ···················· |
| X | 1201 | hCoV-19/England/MILK-BFCCF9/2020\|EPI_ISL_720829\|20 | ···················· | ···················· | ···················· | ···················· | ······T············· | ···················· |
| X | 1202 | hCoV-19/England/MILK-BFF731/2020\|EPI_ISL_720830\|20 | ···················· | ···················· | ···················· | ···················· | ···················· | ···················· |
| X | 1203 | hCoV-19/England/QEUH-C03E8F/2020\|EPI_ISL_720831\|20 | ···················· | ···················· | ···················· | ···················· | ···················· | ···················· |
| X | 1204 | hCoV-19/England/MILK-C00620/2020\|EPI_ISL_720832\|20 | ···················· | ···················· | ···················· | ···················· | ······T············· | ···················· |
| X | 1205 | hCoV-19/England/QEUH-C01407/2020\|EPI_ISL_720834\|20 | ···················· | ···················· | ···················· | ···················· | ···················· | ···················· |
| X | 1206 | hCoV-19/England/MILK-BFCC08/2020\|EPI_ISL_720835\|20 | ···················· | ···················· | ···················· | ···················· | ···················· | ···················· |

| | | | | | | | | |
|---|---|---|---|---|---|---|---|---|
| X | 1207 | hCoV-19/England/ALDP-C04B2D/2020\|EPI_ISL_720836\|20 | ···················· | ···················· | ····················· | ···················· | ···················· | ···················· |
| X | 1208 | hCoV-19/England/MILK-BFF959/2020\|EPI_ISL_720838\|20 | ···················· | ···················· | ····················· | ···················· | ···················· | ···················· |
| X | 1209 | hCoV-19/England/MILK-BFD223/2020\|EPI_ISL_720839\|20 | ···················· | ···················· | ····················· | ···················· | ······T············· | ···················· |
| X | 1210 | hCoV-19/England/MILK-BFCCEA/2020\|EPI_ISL_720840\|20 | ···················· | ···················· | ····················· | ···················· | ···················· | ···················· |
| X | 1211 | hCoV-19/England/QEUH-C0122B/2020\|EPI_ISL_720841\|20 | ···················· | ···················· | ····················· | ···················· | ···················· | ···················· |
| X | 1212 | hCoV-19/England/ALDP-C04BA5/2020\|EPI_ISL_720842\|20 | ···················· | ···················· | ····················· | ···················· | ···················· | ···················· |
| X | 1213 | hCoV-19/England/MILK-BFF90E/2020\|EPI_ISL_720843\|20 | ···················· | ···················· | ····················· | ···················· | ···················· | ···················· |
| X | 1214 | hCoV-19/England/ALDP-C04BE1/2020\|EPI_ISL_720846\|20 | ···················· | ···················· | ····················· | ···················· | ···················· | ···················· |
| X | 1215 | hCoV-19/England/ALDP-C04B96/2020\|EPI_ISL_720848\|20 | ···················· | ···················· | ····················· | ···················· | ···················· | ···················· |
| X | 1216 | hCoV-19/England/ALDP-C04F03/2020\|EPI_ISL_720849\|20 | ···················· | ···················· | ····················· | ···················· | ······T············· | ···················· |
| X | 1217 | hCoV-19/England/QEUH-C0130A/2020\|EPI_ISL_720850\|20 | ···················· | ···················· | ····················· | ···················· | ···················· | ···················· |
| X | 1218 | hCoV-19/England/ALDP-C049AB/2020\|EPI_ISL_720853\|20 | ···················· | ···················· | ····················· | ···················· | ···················· | ···················· |
| X | 1219 | hCoV-19/England/MILK-BFF6CB/2020\|EPI_ISL_720854\|20 | ···················· | ···················· | ····················· | ···················· | ···················· | ···················· |
| X | 1220 | hCoV-19/England/MILK-C006A8/2020\|EPI_ISL_720855\|20 | ···················· | ···················· | ····················· | ···················· | ···················· | ···················· |
| X | 1221 | hCoV-19/England/ALDP-C05C92/2020\|EPI_ISL_720857\|20 | ···················· | ···················· | ····················· | ···················· | ···················· | ···················· |
| X | 1222 | hCoV-19/England/ALDP-C05C65/2020\|EPI_ISL_720858\|20 | ···················· | ···················· | ····················· | ···················· | ···················· | ···················· |
| X | 1223 | hCoV-19/England/MILK-BFCD23/2020\|EPI_ISL_720859\|20 | ···················· | ···················· | ····················· | ···················· | ···················· | ···················· |
| X | 1224 | hCoV-19/England/MILK-BFCF69/2020\|EPI_ISL_720860\|20 | ···················· | ···················· | ····················· | ···················· | ···················· | ···················· |
| X | 1225 | hCoV-19/England/MILK-BFF92C/2020\|EPI_ISL_720861\|20 | ···················· | ···················· | ····················· | ···················· | ···················· | ···················· |
| X | 1226 | hCoV-19/England/MILK-BFCF5A/2020\|EPI_ISL_720862\|20 | ···················· | ···················· | ····················· | ···················· | ···················· | ···················· |
| X | 1227 | hCoV-19/England/ALDP-C05D53/2020\|EPI_ISL_720864\|20 | ···················· | ···················· | ····················· | ···················· | ···················· | ···················· |
| X | 1228 | hCoV-19/England/MILK-BFCBA1/2020\|EPI_ISL_720865\|20 | ···················· | ·A·················· | ····················· | ···················· | ···················· | ··A················· |
| X | 1229 | hCoV-19/England/MILK-C006E4/2020\|EPI_ISL_720868\|20 | ···················· | ···················· | ····················· | ···················· | ···················· | ···················· |
| X | 1230 | hCoV-19/England/ALDP-C04B69/2020\|EPI_ISL_720869\|20 | ···················· | ···················· | ····················· | ···················· | ···················· | ···················· |
| X | 1231 | hCoV-19/England/QEUH-C012C1/2020\|EPI_ISL_720870\|20 | ···················· | ···················· | ····················· | ···················· | ······T············· | ···················· |
| X | 1232 | hCoV-19/England/MILK-BFCF2D/2020\|EPI_ISL_720871\|20 | ···················· | ···················· | ····················· | ···················· | ···················· | ···················· |
| X | 1233 | hCoV-19/England/MILK-BFCE3F/2020\|EPI_ISL_720874\|20 | ···················· | ···················· | ····················· | ···················· | ···················· | ···················· |
| X | 1234 | hCoV-19/England/MILK-C0066C/2020\|EPI_ISL_720876\|20 | ···················· | ···················· | ····················· | ···················· | ···················· | ···················· |
| X | 1235 | hCoV-19/England/QEUH-C013A0/2020\|EPI_ISL_720877\|20 | ···················· | ···················· | ····················· | ···················· | ···················· | ···················· |
| X | 1236 | hCoV-19/England/MILK-C006F3/2020\|EPI_ISL_720878\|20 | ···················· | ···················· | ····················· | ···················· | ···················· | ···················· |
| X | 1237 | hCoV-19/England/MILK-BFF704/2020\|EPI_ISL_720879\|20 | ···················· | ···················· | ····················· | ···················· | ···················· | ···················· |
| X | 1238 | hCoV-19/England/ALDP-C05C74/2020\|EPI_ISL_720880\|20 | ···················· | ···················· | ····················· | ···················· | ···················· | ···················· |
| X | 1239 | hCoV-19/England/MILK-C00611/2020\|EPI_ISL_720882\|20 | ···················· | ···················· | ····················· | ···················· | ···················· | ···················· |
| X | 1240 | hCoV-19/England/MILK-BFF740/2020\|EPI_ISL_720884\|20 | ···················· | ···················· | ····················· | ···················· | ···················· | ···················· |
| X | 1241 | hCoV-19/England/ALDP-C04A5D/2020\|EPI_ISL_720885\|20 | ···················· | ···················· | ····················· | ···················· | ···················· | ···················· |
| X | 1242 | hCoV-19/England/ALDP-C04B4B/2020\|EPI_ISL_720886\|20 | ···················· | ···················· | ····················· | ···················· | ···················· | ···················· |
| X | 1243 | hCoV-19/England/QEUH-C03EAD/2020\|EPI_ISL_720889\|20 | ···················· | ···················· | ····················· | ···················· | ···················· | ···················· |
| X | 1244 | hCoV-19/England/MILK-BFF4B2/2020\|EPI_ISL_720890\|20 | ···················· | ···················· | ····················· | ···················· | ···················· | ···················· |
| X | 1245 | hCoV-19/England/MILK-BFCD32/2020\|EPI_ISL_720892\|20 | ···················· | ···················· | ····················· | ···················· | ···················· | ···················· |
| X | 1246 | hCoV-19/England/ALDP-C04A11/2020\|EPI_ISL_720894\|20 | ···················· | ···················· | ····················· | ···················· | ···················· | ···················· |
| X | 1247 | hCoV-19/England/MILK-C00602/2020\|EPI_ISL_720895\|20 | ···················· | ···················· | ····················· | ···················· | ···················· | ···················· |
| X | 1248 | hCoV-19/England/MILK-BFD205/2020\|EPI_ISL_720896\|20 | ···················· | ···················· | ····················· | ···················· | ···················· | ···················· |
| X | 1249 | hCoV-19/England/QEUH-C03EDA/2020\|EPI_ISL_720897\|20 | ···················· | ···················· | ····················· | ···················· | ···················· | ···················· |
| X | 1250 | hCoV-19/England/ALDP-C05C83/2020\|EPI_ISL_720898\|20 | ···················· | ···················· | ····················· | ···················· | ···················· | ···················· |
| X | 1251 | hCoV-19/England/MILK-BFF607/2020\|EPI_ISL_720899\|20 | ···················· | ···················· | ····················· | ···················· | ···················· | ···················· |
| X | 1252 | hCoV-19/Germany/BW-UKT-a087/2020\|EPI_ISL_723077\|20 | ···················· | ···················· | ····················· | ···················· | ···················· | ···················· |
| X | 1253 | hCoV-19/Germany/BW-UKT-a030/2020\|EPI_ISL_723079\|20 | ···················· | ···················· | ····················· | ···················· | ···················· | ···················· |
| X | 1254 | hCoV-19/Germany/BW-UKT-a021/2020\|EPI_ISL_723080\|20 | ···················· | ···················· | ····················· | ···················· | ···················· | ···················· |
| X | 1255 | hCoV-19/Germany/BW-UKT-a092/2020\|EPI_ISL_723081\|20 | ···················· | ···················· | ····················· | ···················· | ···················· | ···················· |
| X | 1256 | hCoV-19/Germany/BW-UKT-a025/2020\|EPI_ISL_723082\|20 | ···················· | ···················· | ····················· | ···················· | ···················· | ···················· |
| X | 1257 | hCoV-19/Germany/BW-UKT-a112/2020\|EPI_ISL_723083\|20 | ···················· | ···················· | ····················· | ···················· | ···················· | ···················· |
| X | 1258 | hCoV-19/Germany/BW-UKT-a003/2020\|EPI_ISL_723084\|20 | ···················· | ···················· | ····················· | ···················· | ···················· | ···················· |
| X | 1259 | hCoV-19/Germany/BW-UKT-a031/2020\|EPI_ISL_723085\|20 | ···················· | ···················· | ····················· | ···················· | ···················· | ···················· |
| X | 1260 | hCoV-19/Germany/BW-UKT-a110/2020\|EPI_ISL_723086\|20 | ···················· | ···················· | ····················· | ···················· | ···················· | ···················· |
| X | 1261 | hCoV-19/Germany/BW-UKT-a032/2020\|EPI_ISL_723087\|20 | ···················· | ···················· | ····················· | ···················· | ···················· | ···················· |
| X | 1262 | hCoV-19/Germany/BW-UKT-a033/2020\|EPI_ISL_723088\|20 | ···················· | ···················· | ····················· | ···················· | ···················· | ···················· |
| X | 1263 | hCoV-19/Germany/BW-UKT-a035/2020\|EPI_ISL_723089\|20 | ···················· | ···················· | ····················· | ···················· | ···················· | ···················· |
| X | 1264 | hCoV-19/Germany/BW-UKT-a118/2020\|EPI_ISL_723090\|20 | ···················· | ···················· | ····················· | ···················· | ···················· | ···················· |
| X | 1265 | hCoV-19/Germany/BW-UKT-a037/2020\|EPI_ISL_723091\|20 | ···················· | ···················· | ····················· | ···················· | ···················· | ···················· |
| X | 1266 | hCoV-19/Germany/BW-UKT-a038/2020\|EPI_ISL_723092\|20 | ···················· | ···················· | ····················· | ···················· | ···················· | ···················· |
| X | 1267 | hCoV-19/Germany/BW-UKT-a002/2020\|EPI_ISL_723093\|20 | ···················· | ···················· | ····················· | ···················· | ···················· | ···················· |
| X | 1268 | hCoV-19/Germany/BW-UKT-a039/2020\|EPI_ISL_723094\|20 | ···················· | ···················· | ····················· | ···················· | ···················· | ···················· |
| X | 1269 | hCoV-19/Germany/BW-UKT-b003/2020\|EPI_ISL_723095\|20 | ···················· | ···················· | ····················· | ···················· | ···················· | ···················· |
| X | 1270 | hCoV-19/Germany/BW-UKT-a007/2020\|EPI_ISL_723096\|20 | ···················· | ···················· | ····················· | ···················· | ···················· | ···················· |
| X | 1271 | hCoV-19/Germany/BW-UKT-a043/2020\|EPI_ISL_723097\|20 | ···················· | ···················· | ····················· | ···················· | ···················· | ···················· |
| X | 1272 | hCoV-19/Germany/BW-UKT-a005/2020\|EPI_ISL_723098\|20 | ···················· | ···················· | ····················· | ···················· | ···················· | ···················· |
| X | 1273 | hCoV-19/Germany/BW-UKT-a044/2020\|EPI_ISL_723099\|20 | ···················· | ···················· | ····················· | ···················· | ···················· | ···················· |
| X | 1274 | hCoV-19/England/LIVE-DCB22C/2020\|EPI_ISL_724394\|20 | ···················· | ···················· | ····················· | ···················· | ···················· | ···················· |
| X | 1275 | hCoV-19/England/LIVE-DCB550/2020\|EPI_ISL_724410\|20 | ···················· | ···················· | ····················· | ···················· | ···················· | ···················· |
| X | 1276 | hCoV-19/England/LIVE-DCB5AB/2020\|EPI_ISL_724412\|20 | ···················· | ···················· | ····················· | ···················· | ···················· | ···················· |
| X | 1277 | hCoV-19/England/LIVE-DCB5C9/2020\|EPI_ISL_724413\|20 | ···················· | ···················· | ····················· | ···················· | ···················· | ···················· |
| X | 1278 | hCoV-19/England/LIVE-DCB602/2020\|EPI_ISL_724415\|20 | ···················· | ···················· | ····················· | ···················· | ···················· | ···················· |
| X | 1279 | hCoV-19/England/LIVE-DCB6B7/2020\|EPI_ISL_724418\|20 | ···················· | ···················· | ····················· | ···················· | ···················· | ···················· |
| X | 1280 | hCoV-19/England/LIVE-DCB7B4/2020\|EPI_ISL_724427\|20 | ···················· | ···················· | ····················· | ···················· | ···················· | ···················· |
| X | 1281 | hCoV-19/Denmark/DCGC-13813/2020\|EPI_ISL_712410\|202 | ···················· | ···················· | ····················· | ···················· | ···················· | ···················· |
| X | 1282 | hCoV-19/Denmark/DCGC-13816/2020\|EPI_ISL_712411\|202 | ···················· | ···················· | ····················· | ···················· | ···················· | ···················· |
| X | 1283 | hCoV-19/Denmark/DCGC-13826/2020\|EPI_ISL_712412\|202 | ···················· | ···················· | ····················· | ···················· | ···················· | ···················· |
| X | 1284 | hCoV-19/Denmark/DCGC-13829/2020\|EPI_ISL_712413\|202 | ···················· | ···················· | ····················· | ···················· | ···················· | ···················· |
| X | 1285 | hCoV-19/Denmark/DCGC-13839/2020\|EPI_ISL_712414\|202 | ···················· | ···················· | ····················· | ···················· | ···················· | ···················· |
| X | 1286 | hCoV-19/Denmark/DCGC-13886/2020\|EPI_ISL_712416\|202 | ···················· | ···················· | ····················· | ···················· | ···················· | ···················· |

| | | | | | | | | |
|---|---|---|---|---|---|---|---|---|
| X | 1287 | hCoV-19/Denmark/DCGC-13308/2020\|EPI_ISL_712420\|202 | ···················· | ···················· | ···················· | ···················· | ···················· | ···················· |
| X | 1288 | hCoV-19/Denmark/DCGC-13548/2020\|EPI_ISL_712421\|202 | ···················· | ···················· | ···················· | ···················· | ···················· | ···················· |
| X | 1289 | hCoV-19/Denmark/DCGC-13662/2020\|EPI_ISL_712422\|202 | ···················· | ·A·················· | ···················· | ···················· | ···················· | ···A················ |
| X | 1290 | hCoV-19/Denmark/DCGC-13734/2020\|EPI_ISL_712423\|202 | ···················· | ···················· | ···················· | ···················· | ···················· | ···················· |
| X | 1291 | hCoV-19/Denmark/DCGC-13794/2020\|EPI_ISL_712424\|202 | ···················· | ···················· | ···················· | ···················· | ···················· | ···················· |
| X | 1292 | hCoV-19/Denmark/DCGC-13797/2020\|EPI_ISL_712425\|202 | ···················· | ·A·················· | ···················· | ···················· | ···················· | ···A················ |
| X | 1293 | hCoV-19/Denmark/DCGC-13891/2020\|EPI_ISL_712426\|202 | ···················· | ···················· | ···················· | ···················· | ···················· | ···················· |
| X | 1294 | hCoV-19/Denmark/DCGC-12909/2020\|EPI_ISL_712455\|202 | ···················· | ···················· | ···················· | ···················· | ···················· | ···················· |
| X | 1295 | hCoV-19/Denmark/DCGC-12910/2020\|EPI_ISL_712456\|202 | ···················· | ···················· | ···················· | ···················· | ···················· | ···················· |
| X | 1296 | hCoV-19/Denmark/DCGC-12911/2020\|EPI_ISL_712457\|202 | ···················· | ·A·················· | ···················· | ···················· | ···················· | ···················· |
| X | 1297 | hCoV-19/Denmark/DCGC-12912/2020\|EPI_ISL_712458\|202 | ···················· | ···················· | ···················· | ···················· | ···················· | ···················· |
| X | 1298 | hCoV-19/Denmark/DCGC-12914/2020\|EPI_ISL_712459\|202 | ···················· | ···················· | ···················· | ···················· | ···················· | ···················· |
| X | 1299 | hCoV-19/Denmark/DCGC-12916/2020\|EPI_ISL_712460\|202 | ···················· | ···················· | ···················· | ···················· | ···················· | ···················· |
| X | 1300 | hCoV-19/Denmark/DCGC-12917/2020\|EPI_ISL_712461\|202 | ···················· | ···················· | ···················· | ···················· | ···················· | ···················· |
| X | 1301 | hCoV-19/Denmark/DCGC-12918/2020\|EPI_ISL_712462\|202 | ···················· | ···················· | ···················· | ···················· | ···················· | ···················· |
| X | 1302 | hCoV-19/Denmark/DCGC-12919/2020\|EPI_ISL_712463\|202 | ···················· | ···················· | ···················· | ···················· | ···················· | ···················· |
| X | 1303 | hCoV-19/Denmark/DCGC-12921/2020\|EPI_ISL_712464\|202 | ···················· | ···················· | ···················· | ···················· | ···················· | ···················· |
| X | 1304 | hCoV-19/Denmark/DCGC-12922/2020\|EPI_ISL_712465\|202 | ···················· | ···················· | ···················· | ···················· | ···················· | ···················· |
| X | 1305 | hCoV-19/Denmark/DCGC-12923/2020\|EPI_ISL_712466\|202 | ···················· | ···················· | ···················· | ···················· | ···················· | ···················· |
| X | 1306 | hCoV-19/Denmark/DCGC-12924/2020\|EPI_ISL_712467\|202 | ···················· | ···················· | ···················· | ···················· | ···················· | ···················· |
| X | 1307 | hCoV-19/Denmark/DCGC-12927/2020\|EPI_ISL_712468\|202 | ···················· | ···················· | ···················· | ···················· | ···················· | ···················· |
| X | 1308 | hCoV-19/Denmark/DCGC-12930/2020\|EPI_ISL_712469\|202 | ···················· | ···················· | ···················· | ···················· | ···················· | ···················· |
| X | 1309 | hCoV-19/Denmark/DCGC-12931/2020\|EPI_ISL_712470\|202 | ···················· | ···················· | ···················· | ···················· | ···················· | ···················· |
| X | 1310 | hCoV-19/Denmark/DCGC-12932/2020\|EPI_ISL_712471\|202 | ···················· | ···················· | ···················· | ···················· | ···················· | ···················· |
| X | 1311 | hCoV-19/Denmark/DCGC-12933/2020\|EPI_ISL_712472\|202 | ···················· | ·A·················· | ···················· | ···················· | ···················· | ···················· |
| X | 1312 | hCoV-19/Denmark/DCGC-12934/2020\|EPI_ISL_712473\|202 | ···················· | ···················· | ···················· | ···················· | ···················· | ···················· |
| X | 1313 | hCoV-19/Denmark/DCGC-12935/2020\|EPI_ISL_712474\|202 | ···················· | ···················· | ···················· | ···················· | ···················· | ···················· |
| X | 1314 | hCoV-19/Denmark/DCGC-12936/2020\|EPI_ISL_712475\|202 | ···················· | ···················· | ···················· | ···················· | ···················· | ···················· |
| X | 1315 | hCoV-19/Denmark/DCGC-12937/2020\|EPI_ISL_712476\|202 | ···················· | ···················· | ···················· | ···················· | ···················· | ···················· |
| X | 1316 | hCoV-19/Denmark/DCGC-12940/2020\|EPI_ISL_712477\|202 | ···················· | ···················· | ···················· | ···················· | ···················· | ···················· |
| X | 1317 | hCoV-19/Denmark/DCGC-12941/2020\|EPI_ISL_712478\|202 | ···················· | ···················· | ···················· | ···················· | ···················· | ···················· |
| X | 1318 | hCoV-19/Denmark/DCGC-12943/2020\|EPI_ISL_712479\|202 | ···················· | ···················· | ···················· | ···················· | ···················· | ···················· |
| X | 1319 | hCoV-19/Denmark/DCGC-12947/2020\|EPI_ISL_712480\|202 | ···················· | ···················· | ···················· | ···················· | ···················· | ···················· |
| X | 1320 | hCoV-19/Denmark/DCGC-12950/2020\|EPI_ISL_712481\|202 | ···················· | ···················· | ···················· | ···················· | ···················· | ···················· |
| X | 1321 | hCoV-19/Denmark/DCGC-12951/2020\|EPI_ISL_712482\|202 | ···················· | ···················· | ···················· | ···················· | ···················· | ···················· |
| X | 1322 | hCoV-19/Denmark/DCGC-12953/2020\|EPI_ISL_712483\|202 | ···················· | ···················· | ···················· | ···················· | ···················· | ···················· |
| X | 1323 | hCoV-19/Denmark/DCGC-12954/2020\|EPI_ISL_712484\|202 | ···················· | ···················· | ···················· | ···················· | ···················· | ···················· |
| X | 1324 | hCoV-19/Denmark/DCGC-12955/2020\|EPI_ISL_712485\|202 | ···················· | ···················· | ···················· | ···················· | ···················· | ···················· |
| X | 1325 | hCoV-19/Denmark/DCGC-12956/2020\|EPI_ISL_712486\|202 | ···················· | ·A·················· | ···················· | ···················· | ···················· | ···A················ |
| X | 1326 | hCoV-19/Denmark/DCGC-12957/2020\|EPI_ISL_712487\|202 | ···················· | ···················· | ···················· | ···················· | ···················· | ···················· |
| X | 1327 | hCoV-19/Denmark/DCGC-12958/2020\|EPI_ISL_712488\|202 | ···················· | ···················· | ···················· | ···················· | ···················· | ···················· |
| X | 1328 | hCoV-19/Denmark/DCGC-12959/2020\|EPI_ISL_712489\|202 | ···················· | ···················· | ···················· | ···················· | ···················· | ···················· |
| X | 1329 | hCoV-19/Denmark/DCGC-12961/2020\|EPI_ISL_712490\|202 | ···················· | ···················· | ···················· | ···················· | ···················· | ···················· |
| X | 1330 | hCoV-19/Denmark/DCGC-12962/2020\|EPI_ISL_712491\|202 | ···················· | ···················· | ···················· | ···················· | ···················· | ···················· |
| X | 1331 | hCoV-19/Denmark/DCGC-12964/2020\|EPI_ISL_712492\|202 | ···················· | ·A·················· | ···················· | ···················· | ···················· | ···A················ |
| X | 1332 | hCoV-19/Denmark/DCGC-12966/2020\|EPI_ISL_712493\|202 | ···················· | ···················· | ···················· | ···················· | ···················· | ···················· |
| X | 1333 | hCoV-19/Denmark/DCGC-12967/2020\|EPI_ISL_712494\|202 | ···················· | ···················· | ···················· | ···················· | ···················· | ···················· |
| X | 1334 | hCoV-19/Denmark/DCGC-12968/2020\|EPI_ISL_712495\|202 | ···················· | ···················· | ···················· | ···················· | ···················· | ···················· |
| X | 1335 | hCoV-19/Denmark/DCGC-12969/2020\|EPI_ISL_712496\|202 | ···················· | ···················· | ···················· | ···················· | ···················· | ···················· |
| X | 1336 | hCoV-19/Denmark/DCGC-12971/2020\|EPI_ISL_712497\|202 | ···················· | ·············A······ | ···················· | ···················· | ···················· | ···················· |
| X | 1337 | hCoV-19/Denmark/DCGC-12972/2020\|EPI_ISL_712498\|202 | ···················· | ···················· | ···················· | ···················· | ···················· | ···················· |
| X | 1338 | hCoV-19/Denmark/DCGC-12973/2020\|EPI_ISL_712499\|202 | ···················· | ···················· | ···················· | ···················· | ···················· | ···················· |
| X | 1339 | hCoV-19/Denmark/DCGC-15280/2020\|EPI_ISL_713740\|202 | ···················· | ···················· | ···················· | ···················· | ···················· | ···················· |
| X | 1340 | hCoV-19/Denmark/DCGC-15283/2020\|EPI_ISL_713741\|202 | ···················· | ···················· | ···················· | ···················· | ···················· | ···················· |
| X | 1341 | hCoV-19/Denmark/DCGC-15284/2020\|EPI_ISL_713742\|202 | ···················· | ···················· | ···················· | ···················· | ···················· | ···················· |
| X | 1342 | hCoV-19/Denmark/DCGC-15285/2020\|EPI_ISL_713743\|202 | ···················· | ···················· | ···················· | ···················· | ···················· | ···················· |
| X | 1343 | hCoV-19/Denmark/DCGC-15289/2020\|EPI_ISL_713744\|202 | ···················· | ····A··············· | ···················· | ···················· | ···················· | ···················· |
| X | 1344 | hCoV-19/Denmark/DCGC-15290/2020\|EPI_ISL_713745\|202 | ···················· | ···················· | ···················· | ···················· | ···················· | ···················· |
| X | 1345 | hCoV-19/Denmark/DCGC-15296/2020\|EPI_ISL_713748\|202 | ···················· | ···················· | ···················· | ···················· | ···················· | ···················· |
| X | 1346 | hCoV-19/Denmark/DCGC-15297/2020\|EPI_ISL_713749\|202 | ···················· | ···················· | ···················· | ···················· | ···················· | ···················· |
| X | 1347 | hCoV-19/Denmark/DCGC-15298/2020\|EPI_ISL_713750\|202 | ···················· | ···················· | ···················· | ···················· | ···················· | ···················· |
| X | 1348 | hCoV-19/Denmark/DCGC-15299/2020\|EPI_ISL_713751\|202 | ···················· | ···················· | ···················· | ···················· | ···················· | ···················· |
| X | 1349 | hCoV-19/Denmark/DCGC-15300/2020\|EPI_ISL_713752\|202 | ···················· | ···················· | ···················· | ···················· | ···················· | ···················· |
| X | 1350 | hCoV-19/Denmark/DCGC-15304/2020\|EPI_ISL_713753\|202 | ···················· | ···················· | ···················· | ···················· | ···················· | ···················· |
| X | 1351 | hCoV-19/Denmark/DCGC-15306/2020\|EPI_ISL_713754\|202 | ···················· | ···················· | ···················· | ···················· | ···················· | ···················· |
| X | 1352 | hCoV-19/Denmark/DCGC-15307/2020\|EPI_ISL_713755\|202 | ···················· | ···················· | ···················· | ···················· | ···················· | ···················· |
| X | 1353 | hCoV-19/Denmark/DCGC-15310/2020\|EPI_ISL_713756\|202 | ···················· | ···················· | ···················· | ···················· | ···················· | ···················· |
| X | 1354 | hCoV-19/Denmark/DCGC-15311/2020\|EPI_ISL_713757\|202 | ···················· | ···················· | ···················· | ···················· | ···················· | ···················· |
| X | 1355 | hCoV-19/Denmark/DCGC-15312/2020\|EPI_ISL_713758\|202 | ···················· | ···················· | ···················· | ···················· | ···················· | ···················· |
| X | 1356 | hCoV-19/Denmark/DCGC-15353/2020\|EPI_ISL_713780\|202 | ···················· | ···················· | ···················· | ···················· | ···················· | ···················· |
| X | 1357 | hCoV-19/Denmark/DCGC-15354/2020\|EPI_ISL_713781\|202 | ···················· | ···················· | ···················· | ···················· | ···················· | ···················· |
| X | 1358 | hCoV-19/Denmark/DCGC-15356/2020\|EPI_ISL_713782\|202 | ···················· | ·A·················· | ···················· | ···················· | ···················· | ···················· |
| X | 1359 | hCoV-19/Denmark/DCGC-15358/2020\|EPI_ISL_713783\|202 | ···················· | ···················· | ···················· | ···················· | ···················· | ···················· |
| X | 1360 | hCoV-19/Denmark/DCGC-15362/2020\|EPI_ISL_713784\|202 | ···················· | ···················· | ···················· | ···················· | ···················· | ···················· |
| X | 1361 | hCoV-19/Denmark/DCGC-15366/2020\|EPI_ISL_713785\|202 | ···················· | ···················· | ···················· | ···················· | ···················· | ···················· |
| X | 1362 | hCoV-19/Denmark/DCGC-15367/2020\|EPI_ISL_713786\|202 | ···················· | ···················· | ···················· | ···················· | ···················· | ···················· |
| X | 1363 | hCoV-19/Denmark/DCGC-15368/2020\|EPI_ISL_713787\|202 | ···················· | ···················· | ···················· | ···················· | ···················· | ···················· |
| X | 1364 | hCoV-19/Denmark/DCGC-15373/2020\|EPI_ISL_713788\|202 | ···················· | ···················· | ···················· | ···················· | ···················· | ···················· |
| X | 1365 | hCoV-19/Denmark/DCGC-15376/2020\|EPI_ISL_713789\|202 | ···················· | ···················· | ···················· | ···················· | ······T············· | ···················· |
| X | 1366 | hCoV-19/Denmark/DCGC-15377/2020\|EPI_ISL_713790\|202 | ···················· | ···················· | ···················· | ···················· | ···················· | ···················· |

| | | | | | | | | |
|---|---|---|---|---|---|---|---|---|
| X | 1367 | hCoV-19/Denmark/DCGC-15378/2020\|EPI_ISL_713791\|202 | ···················· | ···················· | ···················· | ···················· | ···················· | ···················· |
| X | 1368 | hCoV-19/Denmark/DCGC-15379/2020\|EPI_ISL_713792\|202 | ···················· | ···················· | ···················· | ···················· | ···················· | ···················· |
| X | 1369 | hCoV-19/Denmark/DCGC-15383/2020\|EPI_ISL_713793\|202 | ···················· | ···················· | ···················· | ···················· | ···················· | ···················· |
| X | 1370 | hCoV-19/Denmark/DCGC-15384/2020\|EPI_ISL_713794\|202 | ···················· | ···················· | ···················· | ···················· | ···················· | ···················· |
| X | 1371 | hCoV-19/Denmark/DCGC-15385/2020\|EPI_ISL_713795\|202 | ···················· | ···················· | ···················· | ···················· | ···················· | ···················· |
| X | 1372 | hCoV-19/Denmark/DCGC-15386/2020\|EPI_ISL_713796\|202 | ···················· | ···················· | ···················· | ···················· | ···················· | ···················· |
| X | 1373 | hCoV-19/Denmark/DCGC-15387/2020\|EPI_ISL_713797\|202 | ···················· | ···················· | ···················· | ···················· | ···················· | ···················· |
| X | 1374 | hCoV-19/Denmark/DCGC-15388/2020\|EPI_ISL_713798\|202 | ···················· | ···················· | ···················· | ···················· | ···················· | ···················· |
| X | 1375 | hCoV-19/Denmark/DCGC-15389/2020\|EPI_ISL_713799\|202 | ···················· | ···················· | ···················· | ···················· | ···················· | ···················· |
| X | 1376 | hCoV-19/England/CAMC-C3F6CE/2020\|EPI_ISL_718612\|20 | ···················· | ···················· | ···················· | ···················· | ···················· | ···················· |
| X | 1377 | hCoV-19/England/CAMC-C404A2/2020\|EPI_ISL_718619\|20 | ···················· | ···················· | ···················· | ···················· | ···················· | ···················· |
| X | 1378 | hCoV-19/England/CAMC-C3FD9C/2020\|EPI_ISL_718624\|20 | ···················· | ···················· | ···················· | ···················· | ···················· | ···················· |
| X | 1379 | hCoV-19/England/CAMC-C44D85/2020\|EPI_ISL_718625\|20 | ···················· | ···················· | ···················· | ···················· | ···················· | ···················· |
| X | 1380 | hCoV-19/England/CAMC-C47A21/2020\|EPI_ISL_718626\|20 | ···················· | ···················· | ···················· | ···················· | ···················· | ···················· |
| X | 1381 | hCoV-19/England/ALDP-C498F8/2020\|EPI_ISL_718629\|20 | ···················· | ···················· | ···················· | ···················· | ···················· | ···················· |
| X | 1382 | hCoV-19/England/QEUH-B9EA68/2020\|EPI_ISL_719900\|20 | ···················· | ···················· | ···················· | ···················· | ···················· | ···················· |
| X | 1383 | hCoV-19/England/QEUH-BA2041/2020\|EPI_ISL_719901\|20 | ···················· | ···················· | ···················· | ···················· | ···················· | ···················· |
| X | 1384 | hCoV-19/England/QEUH-B9F171/2020\|EPI_ISL_719902\|20 | ···················· | ···················· | ···················· | ···················· | ···················· | ···················· |
| X | 1385 | hCoV-19/England/QEUH-B9F056/2020\|EPI_ISL_719904\|20 | ···················· | ···················· | ···················· | ···················· | ·····T·············· | ···················· |
| X | 1386 | hCoV-19/England/QEUH-B9F126/2020\|EPI_ISL_719907\|20 | ···················· | ···················· | ···················· | ···················· | ···················· | ···················· |
| X | 1387 | hCoV-19/England/QEUH-B9F162/2020\|EPI_ISL_719908\|20 | ···················· | ···················· | ···················· | ···················· | ···················· | ···················· |
| X | 1388 | hCoV-19/England/QEUH-B9E813/2020\|EPI_ISL_719909\|20 | ···················· | ···················· | ···················· | ···················· | ···················· | ···················· |
| X | 1389 | hCoV-19/England/QEUH-B9E85F/2020\|EPI_ISL_719910\|20 | ···················· | ···················· | ···················· | ···················· | ···················· | ···················· |
| X | 1390 | hCoV-19/England/QEUH-BA275B/2020\|EPI_ISL_719911\|20 | ···················· | ···················· | ···················· | ···················· | ·····T·············· | ···················· |
| X | 1391 | hCoV-19/England/QEUH-B9F00B/2020\|EPI_ISL_719912\|20 | ···················· | ···················· | ···················· | ···················· | ···················· | ···················· |
| X | 1392 | hCoV-19/England/QEUH-B9E743/2020\|EPI_ISL_719913\|20 | ···················· | ···················· | ···················· | ···················· | ···················· | ···················· |
| X | 1393 | hCoV-19/England/QEUH-B9E716/2020\|EPI_ISL_719914\|20 | ···················· | ···················· | ···················· | ···················· | ···················· | ···················· |
| X | 1394 | hCoV-19/England/QEUH-B9EA59/2020\|EPI_ISL_719915\|20 | ···················· | ···················· | ···················· | ···················· | ···················· | ···················· |
| X | 1395 | hCoV-19/England/QEUH-BA1877/2020\|EPI_ISL_719916\|20 | ···················· | ···················· | ···················· | ···················· | ···················· | ···················· |
| X | 1396 | hCoV-19/England/QEUH-BA22D2/2020\|EPI_ISL_719917\|20 | ···················· | ···················· | ···················· | ···················· | ···················· | ···················· |
| X | 1397 | hCoV-19/England/QEUH-BA18A4/2020\|EPI_ISL_719918\|20 | ···················· | ···················· | ···················· | ···················· | ···················· | ···················· |
| X | 1398 | hCoV-19/England/QEUH-B9EB83/2020\|EPI_ISL_719920\|20 | ···················· | ···················· | ···················· | ···················· | ···················· | ···················· |
| X | 1399 | hCoV-19/England/QEUH-B9E673/2020\|EPI_ISL_719921\|20 | ···················· | ···················· | ···················· | ···················· | ···················· | ···················· |
| X | 1400 | hCoV-19/England/QEUH-B9F0DE/2020\|EPI_ISL_719940\|20 | ···················· | ···················· | ···················· | ···················· | ···················· | ···················· |
| X | 1401 | hCoV-19/England/QEUH-BA2375/2020\|EPI_ISL_719941\|20 | ···················· | ···················· | ···················· | ···················· | ·····T·············· | ···················· |
| X | 1402 | hCoV-19/England/QEUH-BA27E2/2020\|EPI_ISL_719944\|20 | ···················· | ···················· | ···················· | ···················· | ···················· | ···················· |
| X | 1403 | hCoV-19/England/QEUH-BA1FDC/2020\|EPI_ISL_719946\|20 | ···················· | ···················· | ···················· | ···················· | ···················· | ···················· |
| X | 1404 | hCoV-19/England/QEUH-BA27C4/2020\|EPI_ISL_719947\|20 | ···················· | ···················· | ···················· | ···················· | ···················· | ···················· |
| X | 1405 | hCoV-19/England/QEUH-BA1B8D/2020\|EPI_ISL_719951\|20 | ···················· | ···················· | ···················· | ···················· | ···················· | ···················· |
| X | 1406 | hCoV-19/England/QEUH-B9F108/2020\|EPI_ISL_719953\|20 | ···················· | ···················· | ···················· | ···················· | ···················· | ···················· |
| X | 1407 | hCoV-19/England/QEUH-B9EEC6/2020\|EPI_ISL_719956\|20 | ···················· | ···················· | ···················· | ···················· | ···················· | ···················· |
| X | 1408 | hCoV-19/England/QEUH-B9EFF0/2020\|EPI_ISL_719957\|20 | ···················· | ···················· | ···················· | ···················· | ···················· | ···················· |
| X | 1409 | hCoV-19/England/QEUH-B9EAE0/2020\|EPI_ISL_719958\|20 | ···················· | ···················· | ···················· | ···················· | ·····T·············· | ···················· |
| X | 1410 | hCoV-19/England/QEUH-BA2788/2020\|EPI_ISL_719959\|20 | ···················· | ···················· | ···················· | ···················· | ···················· | ···················· |
| X | 1411 | hCoV-19/England/QEUH-BA3286/2020\|EPI_ISL_720920\|20 | ···················· | ···················· | ···················· | ···················· | ···················· | ···················· |
| X | 1412 | hCoV-19/England/QEUH-BA2D4A/2020\|EPI_ISL_720921\|20 | ···················· | ···················· | ···················· | ···················· | ···················· | ···················· |
| X | 1413 | hCoV-19/England/QEUH-B9F724/2020\|EPI_ISL_720922\|20 | ···················· | ···················· | ···················· | ···················· | ···················· | ···················· |
| X | 1414 | hCoV-19/England/QEUH-BA3602/2020\|EPI_ISL_720924\|20 | ···················· | ···················· | ···················· | ···················· | ···················· | ···················· |
| X | 1415 | hCoV-19/England/QEUH-B9E0EE/2020\|EPI_ISL_720926\|20 | ···················· | ···················· | ···················· | ···················· | ···················· | ···················· |
| X | 1416 | hCoV-19/England/QEUH-B9E215/2020\|EPI_ISL_720927\|20 | ···················· | ···················· | ···················· | ···················· | ···················· | ···················· |
| X | 1417 | hCoV-19/England/QEUH-BA41D3/2020\|EPI_ISL_720929\|20 | ···················· | ·A·················· | ···················· | ···················· | ···················· | ··A················· |
| X | 1418 | hCoV-19/England/QEUH-BA35F6/2020\|EPI_ISL_720930\|20 | ···················· | ···················· | ···················· | ···················· | ···················· | ···················· |
| X | 1419 | hCoV-19/England/QEUH-B9FDF5/2020\|EPI_ISL_720931\|20 | ···················· | ···················· | ···················· | ···················· | ···················· | ···················· |
| X | 1420 | hCoV-19/England/QEUH-BA2DD1/2020\|EPI_ISL_720932\|20 | ···················· | ···················· | ···················· | ···················· | ···················· | ···················· |
| X | 1421 | hCoV-19/England/QEUH-B9FDC8/2020\|EPI_ISL_720933\|20 | ···················· | ···················· | ···················· | ···················· | ···················· | ···················· |
| X | 1422 | hCoV-19/England/QEUH-B9F8E5/2020\|EPI_ISL_720934\|20 | ···················· | ···················· | ···················· | ···················· | ···················· | ···················· |
| X | 1423 | hCoV-19/England/QEUH-B9E46A/2020\|EPI_ISL_720936\|20 | ···················· | ···················· | ···················· | ···················· | ···················· | ···················· |
| X | 1424 | hCoV-19/England/QEUH-B9F51B/2020\|EPI_ISL_720937\|20 | ···················· | ···················· | ···················· | ···················· | ···················· | ···················· |
| X | 1425 | hCoV-19/England/QEUH-B9F593/2020\|EPI_ISL_720938\|20 | ···················· | ···················· | ···················· | ···················· | ···················· | ···················· |
| X | 1426 | hCoV-19/England/QEUH-B9FB82/2020\|EPI_ISL_720939\|20 | ···················· | ···················· | ···················· | ···················· | ···················· | ···················· |
| X | 1427 | hCoV-19/England/QEUH-B9E38B/2020\|EPI_ISL_720940\|20 | ···················· | ···················· | ···················· | ···················· | ···················· | ···················· |
| X | 1428 | hCoV-19/England/QEUH-B9F9E2/2020\|EPI_ISL_720941\|20 | ···················· | ···················· | ···················· | ···················· | ···················· | ···················· |
| X | 1429 | hCoV-19/Germany/BW-UKT-a006/2020\|EPI_ISL_723100\|20 | ···················· | ···················· | ···················· | ···················· | ···················· | ···················· |
| X | 1430 | hCoV-19/Germany/BW-UKT-b007/2020\|EPI_ISL_723101\|20 | ···················· | ···················· | ···················· | ···················· | ···················· | ···················· |
| X | 1431 | hCoV-19/Germany/BW-UKT-a009/2020\|EPI_ISL_723102\|20 | ···················· | ···················· | ···················· | ···················· | ···················· | ···················· |
| X | 1432 | hCoV-19/Germany/BW-UKT-a046/2020\|EPI_ISL_723103\|20 | ···················· | ···················· | ···················· | ···················· | ···················· | ···················· |
| X | 1433 | hCoV-19/Germany/BW-UKT-a053/2020\|EPI_ISL_723104\|20 | ···················· | ···················· | ···················· | ···················· | ···················· | ···················· |
| X | 1434 | hCoV-19/Germany/BW-UKT-a013/2020\|EPI_ISL_723105\|20 | ···················· | ···················· | ···················· | ···················· | ···················· | ···················· |
| X | 1435 | hCoV-19/Germany/BW-UKT-a014/2020\|EPI_ISL_723106\|20 | ···················· | ···················· | ···················· | ···················· | ···················· | ···················· |
| X | 1436 | hCoV-19/Germany/BW-UKT-a113/2020\|EPI_ISL_723107\|20 | ···················· | ···················· | ···················· | ···················· | ···················· | ···················· |
| X | 1437 | hCoV-19/Germany/BW-UKT-a016/2020\|EPI_ISL_723108\|20 | ···················· | ···················· | ···················· | ···················· | ···················· | ···················· |
| X | 1438 | hCoV-19/Germany/BW-UKT-a117/2020\|EPI_ISL_723109\|20 | ···················· | ···················· | ···················· | ···················· | ···················· | ···················· |
| X | 1439 | hCoV-19/Germany/BW-UKT-a017/2020\|EPI_ISL_723110\|20 | ···················· | ···················· | ···················· | ···················· | ···················· | ···················· |
| X | 1440 | hCoV-19/Germany/BW-UKT-a020/2020\|EPI_ISL_723111\|20 | ···················· | ···················· | ···················· | ···················· | ···················· | ···················· |
| X | 1441 | hCoV-19/Germany/BW-UKT-a068/2020\|EPI_ISL_723112\|20 | ···················· | ···················· | ···················· | ···················· | ···················· | ···················· |
| X | 1442 | hCoV-19/Germany/BW-UKT-a085/2020\|EPI_ISL_723113\|20 | ···················· | ···················· | ···················· | ···················· | ···················· | ···················· |
| X | 1443 | hCoV-19/Germany/BW-UKT-a086/2020\|EPI_ISL_723114\|20 | ···················· | ···················· | ···················· | ···················· | ···················· | ···················· |
| X | 1444 | hCoV-19/Germany/BW-UKT-a022/2020\|EPI_ISL_723115\|20 | ···················· | ···················· | ···················· | ···················· | ···················· | ···················· |
| X | 1445 | hCoV-19/Germany/BW-UKT-a089/2020\|EPI_ISL_723116\|20 | ···················· | ···················· | ···················· | ···················· | ···················· | ···················· |
| X | 1446 | hCoV-19/Germany/BW-UKT-a090/2020\|EPI_ISL_723117\|20 | ···················· | ···················· | ···················· | ···················· | ···················· | ···················· |

```
X 1447 hCoV-19/USA/MN-MDH-2167/2020|EPI_ISL_723118|2020-0 ···················· ···················· ····················· ···················· ···················· ····················
X 1448 hCoV-19/USA/MN-MDH-2168/2020|EPI_ISL_723119|2020-1 ···················· ···················· ····················· ···················· ···················· ····················
X 1450 hCoV-19/USA/MN-MDH-2170/2020|EPI_ISL_723121|2020-1 ···················· ···················· ····················· ···················· ···················· ····················
X 1451 hCoV-19/USA/MN-MDH-2171/2020|EPI_ISL_723122|2020-1 ···················· ···················· ····················· ···················· ···················· ····················
X 1452 hCoV-19/USA/MN-MDH-2172/2020|EPI_ISL_723123|2020-1 ···················· ···················· ····················· ···················· ···················· ····················
X 1453 hCoV-19/USA/MN-MDH-2173/2020|EPI_ISL_723124|2020-1 ···················· ···················· ····················· ···················· ···················· ····················
X 1454 hCoV-19/USA/MN-MDH-2174/2020|EPI_ISL_723125|2020-1 ···················· ···················· ····················· ···················· ···················· ····················
X 1455 hCoV-19/USA/MN-MDH-2175/2020|EPI_ISL_723126|2020-1 ···················· ···················· ····················· ···················· ···················· ····················
X 1456 hCoV-19/USA/MN-MDH-2176/2020|EPI_ISL_723127|2020-1 ···················· ···················· ····················· ···················· ···················· ····················
X 1457 hCoV-19/USA/MN-MDH-2177/2020|EPI_ISL_723128|2020-1 ···················· ···················· ····················· ···················· ···················· ····················
X 1458 hCoV-19/USA/MN-MDH-2178/2020|EPI_ISL_723129|2020-1 ···················· ···················· ····················· ···················· ···················· ····················
X 1459 hCoV-19/USA/MN-MDH-2179/2020|EPI_ISL_723130|2020-1 ···················· ···················· ····················· ···················· ···················· ····················
X 1460 hCoV-19/USA/MN-MDH-2180/2020|EPI_ISL_723131|2020-1 ···················· ···················· ····················· ···················· ···················· ····················
X 1461 hCoV-19/USA/MN-MDH-2181/2020|EPI_ISL_723132|2020-1 ···················· ···················· ····················· ···················· ···················· ····················
X 1462 hCoV-19/USA/MN-MDH-2182/2020|EPI_ISL_723133|2020-1 ···················· ···················· ····················· ···················· ···················· ····················
X 1463 hCoV-19/USA/MN-MDH-2183/2020|EPI_ISL_723134|2020-1 ···················· ···················· ····················· ···················· ···················· ····················
X 1464 hCoV-19/USA/MN-MDH-2184/2020|EPI_ISL_723135|2020-1 ···················· ···················· ····················· ···················· ···················· ····················
X 1465 hCoV-19/USA/MN-MDH-2185/2020|EPI_ISL_723136|2020-1 ···················· ···················· ····················· ···················· ···················· ····················
X 1466 hCoV-19/USA/MN-MDH-2186/2020|EPI_ISL_723137|2020-1 ···················· ···················· ····················· ···················· ···················· ····················
X 1467 hCoV-19/USA/MN-MDH-2187/2020|EPI_ISL_723138|2020-1 ···················· ···················· ····················· ···················· ···················· ····················
X 1468 hCoV-19/USA/MN-MDH-2188/2020|EPI_ISL_723139|2020-1 ···················· ···················· ····················· ···················· ···················· ····················
X 1469 hCoV-19/USA/MN-MDH-2189/2020|EPI_ISL_723140|2020-1 ···················· ···················· ····················· ···················· ···················· ····················
X 1470 hCoV-19/USA/MN-MDH-2190/2020|EPI_ISL_723141|2020-1 ···················· ···················· ····················· ···················· ···················· ····················
X 1471 hCoV-19/USA/MN-MDH-2191/2020|EPI_ISL_723142|2020-1 ···················· ···················· ····················· ···················· ···················· ····················
X 1472 hCoV-19/USA/MN-MDH-2192/2020|EPI_ISL_723143|2020-1 ···················· ···················· ····················· ···················· ···················· ····················
X 1473 hCoV-19/USA/MN-MDH-2193/2020|EPI_ISL_723144|2020-1 ···················· ···················· ····················· ···················· ···················· ····················
X 1474 hCoV-19/USA/MN-MDH-2194/2020|EPI_ISL_723145|2020-1 ···················· ···················· ····················· ···················· ···················· ····················
X 1475 hCoV-19/USA/MN-MDH-2195/2020|EPI_ISL_723146|2020-1 ···················· ···················· ····················· ···················· ···················· ····················
X 1476 hCoV-19/USA/MN-MDH-2196/2020|EPI_ISL_723147|2020-1 ···················· ···················· ····················· ···················· ···················· ····················
X 1477 hCoV-19/USA/MN-MDH-2197/2020|EPI_ISL_723148|2020-1 ···················· ···················· ····················· ···················· ···················· ····················
X 1478 hCoV-19/USA/MN-MDH-2198/2020|EPI_ISL_723149|2020-1 ···················· ···················· ····················· ···················· ···················· ····················
X 1479 hCoV-19/USA/MN-MDH-2199/2020|EPI_ISL_723150|2020-1 ···················· ·A·················· ····················· ···················· ···················· ····················
X 1480 hCoV-19/USA/MN-MDH-2200/2020|EPI_ISL_723151|2020-1 ···················· ···················· ····················· ···················· ···················· ····················
X 1481 hCoV-19/USA/MN-MDH-2201/2020|EPI_ISL_723152|2020-1 ···················· ···················· ····················· ···················· ···················· ····················
X 1482 hCoV-19/USA/MN-MDH-2202/2020|EPI_ISL_723153|2020-1 ···················· ···················· ····················· ···················· ···················· ····················
X 1483 hCoV-19/USA/MN-MDH-2203/2020|EPI_ISL_723154|2020-1 ···················· ···················· ····················· ···················· ···················· ····················
X 1484 hCoV-19/Netherlands/UT-RIVM-20180/2020|EPI_ISL_723 ···················· ···················· ···T················· ···················· ···················· ····················
X 1485 hCoV-19/Netherlands/NH-RIVM-20112/2020|EPI_ISL_723 ···················· ···················· ····················· ···················· ···················· ····················
X 1486 hCoV-19/Netherlands/NB-RIVM-20265/2020|EPI_ISL_723 ···················· ···················· ····················· ···················· ···················· ····················
X 1487 hCoV-19/Netherlands/GE-RIVM-20046/2020|EPI_ISL_723 ···················· ···················· ····················· ···················· ···················· ····················
X 1488 hCoV-19/Netherlands/GE-RIVM-20047/2020|EPI_ISL_723 ···················· ···················· ····················· ···················· ···················· ····················
X 1489 hCoV-19/Netherlands/UT-RIVM-20068/2020|EPI_ISL_723 ···················· ···················· ····················· ···················· ···················· ····················
X 1490 hCoV-19/Netherlands/GE-RIVM-20071/2020|EPI_ISL_723 ···················· ···················· ····················· ···················· ···················· ····················
X 1491 hCoV-19/Netherlands/GE-RIVM-20072/2020|EPI_ISL_723 ···················· ···················· ····················· ···················· ···················· ····················
X 1492 hCoV-19/Netherlands/GE-RIVM-20073/2020|EPI_ISL_723 ···················· ···················· ····················· ···················· ···················· ····················
X 1493 hCoV-19/Netherlands/OV-RIVM-20099/2020|EPI_ISL_723 ···················· ···················· ····················· ···················· ···················· ····················
X 1494 hCoV-19/Netherlands/GE-RIVM-20126/2020|EPI_ISL_723 ···················· ···················· ····················· ···················· ···················· ····················
X 1495 hCoV-19/Netherlands/NH-RIVM-20131/2020|EPI_ISL_723 ···················· ···················· ····················· ···················· ···················· ····················
X 1496 hCoV-19/Netherlands/GE-RIVM-20154/2020|EPI_ISL_723 ···················· ···················· ····················· ···················· ···················· ····················
X 1497 hCoV-19/Netherlands/GE-RIVM-20155/2020|EPI_ISL_723 ···················· ···················· ····················· ···················· ···················· ····················
X 1498 hCoV-19/Netherlands/UT-RIVM-20165/2020|EPI_ISL_723 ···················· ···················· ····················· ···················· ···················· ····················
X 1499 hCoV-19/Netherlands/GE-RIVM-20169/2020|EPI_ISL_723 ···················· ···················· ····················· ···················· ···················· ····················
X 1500 hCoV-19/Netherlands/GE-RIVM-20170/2020|EPI_ISL_723 ···················· ···················· ····················· ···················· ···················· ····················
X 1501 hCoV-19/Netherlands/GE-RIVM-20171/2020|EPI_ISL_723 ···················· ···················· ····················· ···················· ···················· ····················
X 1502 hCoV-19/Netherlands/GE-RIVM-20172/2020|EPI_ISL_723 ···················· ···················· ····················· ···················· ···················· ····················
X 1503 hCoV-19/Netherlands/GE-RIVM-20173/2020|EPI_ISL_723 ···················· ···················· ····················· ···················· ···················· ····················
X 1504 hCoV-19/Netherlands/NH-RIVM-20233/2020|EPI_ISL_723 ···················· ···················· ····················· ···················· ···················· ····················
X 1505 hCoV-19/Netherlands/LI-RIVM-20241/2020|EPI_ISL_723 ···················· ···················· ····················· ···················· ···················· ····················
X 1506 hCoV-19/Netherlands/LI-RIVM-20244/2020|EPI_ISL_723 ···················· ···················· ····················· ···················· ···················· ····················
X 1507 hCoV-19/Netherlands/GE-RIVM-20264/2020|EPI_ISL_723 ···················· ···················· ····················· ···················· ···················· ····················
X 1508 hCoV-19/Netherlands/NB-RIVM-20267/2020|EPI_ISL_723 ···················· ···················· ····················· ···················· ···················· ····················
X 1509 hCoV-19/England/LIVE-DCB80C/2020|EPI_ISL_724430|20 ···················· ·········T·········· ····················· ···················· ···················· ····················
X 1510 hCoV-19/England/LIVE-DCB839/2020|EPI_ISL_724431|20 ···················· ···················· ····················· ···················· ···················· ····················
X 1511 hCoV-19/England/LIVE-DCB866/2020|EPI_ISL_724434|20 ···················· ···················· ····················· ···················· ···················· ····················
X 1512 hCoV-19/England/LIVE-DCB8B1/2020|EPI_ISL_724436|20 ···················· ···················· ····················· ···················· ···················· ····················
X 1513 hCoV-19/England/LIVE-DCB909/2020|EPI_ISL_724438|20 ···················· ···················· ····················· ···················· ···················· ····················
X 1514 hCoV-19/England/LIVE-DCB972/2020|EPI_ISL_724442|20 ···················· ···················· ····················· ···················· ···················· ····················
X 1515 hCoV-19/England/LIVE-DCB981/2020|EPI_ISL_724443|20 ···················· ···················· ····················· ···················· ···················· ····················
X 1516 hCoV-19/England/LIVE-DCB9AF/2020|EPI_ISL_724444|20 ···················· ···················· ····················· ···················· ···················· ····················
X 1517 hCoV-19/England/LIVE-DCBA24/2020|EPI_ISL_724446|20 ···················· ···················· ····················· ···················· ···················· ····················
X 1518 hCoV-19/England/LIVE-DCBA51/2020|EPI_ISL_724448|20 ···················· ···················· ····················· ···················· ······T············· ····················
X 1519 hCoV-19/England/LIVE-DCBA7F/2020|EPI_ISL_724449|20 ···················· ···················· ····················· ···················· ···················· ····················
X 1520 hCoV-19/England/LIVE-DCBB6D/2020|EPI_ISL_724451|20 ···················· ···················· ····················· ···················· ···················· ····················
X 1521 hCoV-19/England/LIVE-DCBC2E/2020|EPI_ISL_724452|20 ···················· ···················· ····················· ···················· ···················· ····················
X 1522 hCoV-19/England/LIVE-DCBC4C/2020|EPI_ISL_724454|20 ···················· ···················· ····················· ···················· ···················· ····················
X 1523 hCoV-19/England/LIVE-DCBD0D/2020|EPI_ISL_724458|20 ···················· ···················· ····················· ···················· ···················· ····················
X 1524 hCoV-19/England/LIVE-DCBD2B/2020|EPI_ISL_724460|20 ···················· ···················· ····················· ···················· ···················· ····················
X 1525 hCoV-19/England/LIVE-DCBDA3/2020|EPI_ISL_724463|20 ···················· ···················· ····················· ···················· ···················· ····················
X 1526 hCoV-19/England/LIVE-DCBDB2/2020|EPI_ISL_724464|20 ···················· ···················· ····················· ···················· ···················· ····················
X 1527 hCoV-19/England/LIVE-DCBDD0/2020|EPI_ISL_724465|20 ···················· ···················· ····················· ···················· ···················· ····················
```

| | | | | | | | | |
|---|---|---|---|---|---|---|---|---|
| X | 1528 | hCoV-19/England/LIVE-DCBE28/2020\|EPI_ISL_724467\|20 | ···················· | ···················· | ···················· | ···················· | ···················· | ···················· |
| X | 1529 | hCoV-19/England/LIVE-DCBE37/2020\|EPI_ISL_724468\|20 | ···················· | ···················· | ···················· | ···················· | ···················· | ···················· |
| X | 1530 | hCoV-19/England/LIVE-DCBE91/2020\|EPI_ISL_724472\|20 | ···················· | ···················· | ···················· | ···················· | ···················· | ···················· |
| X | 1531 | hCoV-19/England/LIVE-DCBEBF/2020\|EPI_ISL_724474\|20 | ···················· | ···················· | ···················· | ···················· | ···················· | ···················· |
| X | 1532 | hCoV-19/England/LIVE-DCBFDA/2020\|EPI_ISL_724479\|20 | ·T·················· | ···················· | ···················· | ···················· | ···················· | ···················· |
| X | 1533 | hCoV-19/England/LIVE-DCC021/2020\|EPI_ISL_724481\|20 | ···················· | ···················· | ···················· | ···················· | ···················· | ···················· |
| X | 1534 | hCoV-19/England/LIVE-DCC030/2020\|EPI_ISL_724482\|20 | ···················· | ···················· | ···················· | ···················· | ···················· | ···················· |
| X | 1535 | hCoV-19/England/LIVE-DCC04F/2020\|EPI_ISL_724483\|20 | ···················· | ···················· | ···················· | ···················· | ···················· | ···················· |
| X | 1536 | hCoV-19/England/LIVE-DCC0D6/2020\|EPI_ISL_724487\|20 | ···················· | ···················· | ···················· | ···················· | ···················· | ···················· |
| X | 1537 | hCoV-19/Denmark/DCGC-12974/2020\|EPI_ISL_712500\|202 | ···················· | ···················· | ···················· | ···················· | ·····T·············· | ···················· |
| X | 1538 | hCoV-19/Denmark/DCGC-12975/2020\|EPI_ISL_712501\|202 | ···················· | ···················· | ···················· | ···················· | ···················· | ···················· |
| X | 1539 | hCoV-19/Denmark/DCGC-12977/2020\|EPI_ISL_712502\|202 | ···················· | ···················· | ···················· | ···················· | ···················· | ···················· |
| X | 1540 | hCoV-19/Denmark/DCGC-12978/2020\|EPI_ISL_712503\|202 | ···················· | ···················· | ···················· | ···················· | ···················· | ···················· |
| X | 1541 | hCoV-19/Denmark/DCGC-12980/2020\|EPI_ISL_712504\|202 | ···················· | ·A·················· | ···················· | ···················· | ···················· | ···················· |
| X | 1542 | hCoV-19/Denmark/DCGC-12981/2020\|EPI_ISL_712505\|202 | ···················· | ···················· | ···················· | ···················· | ···················· | ···················· |
| X | 1543 | hCoV-19/Denmark/DCGC-12982/2020\|EPI_ISL_712506\|202 | ···················· | ···················· | ···················· | ···················· | ···················· | ···················· |
| X | 1544 | hCoV-19/Denmark/DCGC-12983/2020\|EPI_ISL_712507\|202 | ···················· | ·A·················· | ···················· | ···················· | ···················· | ···················· |
| X | 1545 | hCoV-19/Denmark/DCGC-12984/2020\|EPI_ISL_712508\|202 | ···················· | ···················· | ···················· | ···················· | ···················· | ···················· |
| X | 1546 | hCoV-19/Denmark/DCGC-12985/2020\|EPI_ISL_712509\|202 | ···················· | ···················· | ···················· | ···················· | ···················· | ···················· |
| X | 1547 | hCoV-19/Denmark/DCGC-12986/2020\|EPI_ISL_712510\|202 | ···················· | ···················· | ···················· | ···················· | ···················· | ···················· |
| X | 1548 | hCoV-19/Denmark/DCGC-12988/2020\|EPI_ISL_712511\|202 | ···················· | ···················· | ···················· | ···················· | ···················· | ···················· |
| X | 1549 | hCoV-19/Denmark/DCGC-12989/2020\|EPI_ISL_712512\|202 | ···················· | ···················· | ···················· | ···················· | ···················· | ···················· |
| X | 1550 | hCoV-19/Denmark/DCGC-12991/2020\|EPI_ISL_712513\|202 | ···················· | ···················· | ···················· | ···················· | ···················· | ···················· |
| X | 1551 | hCoV-19/Denmark/DCGC-12992/2020\|EPI_ISL_712514\|202 | ···················· | ···················· | ···················· | ···················· | ···················· | ···················· |
| X | 1552 | hCoV-19/Denmark/DCGC-12993/2020\|EPI_ISL_712515\|202 | ···················· | ···················· | ···················· | ···················· | ···················· | ···················· |
| X | 1553 | hCoV-19/Denmark/DCGC-12995/2020\|EPI_ISL_712516\|202 | ···················· | ···················· | ···················· | ···················· | ···················· | ···················· |
| X | 1554 | hCoV-19/Denmark/DCGC-12997/2020\|EPI_ISL_712517\|202 | ···················· | ···················· | ···················· | ···················· | ···················· | ···················· |
| X | 1555 | hCoV-19/Denmark/DCGC-12999/2020\|EPI_ISL_712518\|202 | ···················· | ···················· | ···················· | ···················· | ···················· | ···················· |
| X | 1556 | hCoV-19/Denmark/DCGC-13000/2020\|EPI_ISL_712519\|202 | ···················· | ···················· | ···················· | ···················· | ···················· | ···················· |
| X | 1557 | hCoV-19/Denmark/DCGC-13001/2020\|EPI_ISL_712520\|202 | ···················· | ···················· | ···················· | ···················· | ···················· | ···················· |
| X | 1558 | hCoV-19/Denmark/DCGC-13003/2020\|EPI_ISL_712521\|202 | ···················· | ···················· | ···················· | ···················· | ···················· | ···················· |
| X | 1559 | hCoV-19/Denmark/DCGC-13004/2020\|EPI_ISL_712522\|202 | ···················· | ···················· | ···················· | ···················· | ···················· | ···················· |
| X | 1560 | hCoV-19/Denmark/DCGC-13006/2020\|EPI_ISL_712523\|202 | ···················· | ···················· | ···················· | ···················· | ···················· | ···················· |
| X | 1561 | hCoV-19/Denmark/DCGC-13008/2020\|EPI_ISL_712524\|202 | ···················· | ···················· | ···················· | ···················· | ···················· | ···················· |
| X | 1562 | hCoV-19/Denmark/DCGC-13009/2020\|EPI_ISL_712525\|202 | ···················· | ···················· | ···················· | ···················· | ···················· | ···················· |
| X | 1563 | hCoV-19/Denmark/DCGC-13010/2020\|EPI_ISL_712526\|202 | ···················· | ···················· | ···················· | ···················· | ···················· | ···················· |
| X | 1564 | hCoV-19/Denmark/DCGC-13040/2020\|EPI_ISL_712549\|202 | ···················· | ···················· | ···················· | ···················· | ···················· | ···················· |
| X | 1565 | hCoV-19/Denmark/DCGC-13045/2020\|EPI_ISL_712552\|202 | ···················· | ···················· | ···················· | ···················· | ···················· | ···················· |
| X | 1566 | hCoV-19/Denmark/DCGC-13049/2020\|EPI_ISL_712554\|202 | ···················· | ···················· | ···················· | ···················· | ···················· | ···················· |
| X | 1567 | hCoV-19/Denmark/DCGC-13053/2020\|EPI_ISL_712555\|202 | ···················· | ···················· | ···················· | ···················· | ···················· | ···················· |
| X | 1568 | hCoV-19/Denmark/DCGC-13056/2020\|EPI_ISL_712556\|202 | ···················· | ···················· | ···················· | ···················· | ···················· | ···················· |
| X | 1569 | hCoV-19/Denmark/DCGC-13058/2020\|EPI_ISL_712557\|202 | ···················· | ···················· | ···················· | ···················· | ···················· | ···················· |
| X | 1570 | hCoV-19/Denmark/DCGC-13059/2020\|EPI_ISL_712558\|202 | ···················· | ···················· | ···················· | ···················· | ···················· | ···················· |
| X | 1571 | hCoV-19/Denmark/DCGC-13060/2020\|EPI_ISL_712559\|202 | ···················· | ···················· | ···················· | ···················· | ···················· | ···················· |
| X | 1572 | hCoV-19/Denmark/DCGC-15394/2020\|EPI_ISL_713800\|202 | ···················· | ···················· | ···················· | ···················· | ···················· | ···················· |
| X | 1573 | hCoV-19/Denmark/DCGC-15396/2020\|EPI_ISL_713801\|202 | ···················· | ···················· | ···················· | ···················· | ···················· | ···················· |
| X | 1574 | hCoV-19/Denmark/DCGC-15398/2020\|EPI_ISL_713802\|202 | ···················· | ···················· | ···················· | ···················· | ···················· | ···················· |
| X | 1575 | hCoV-19/Denmark/DCGC-15399/2020\|EPI_ISL_713803\|202 | ···················· | ···················· | ···················· | ···················· | ···················· | ···················· |
| X | 1576 | hCoV-19/Denmark/DCGC-15400/2020\|EPI_ISL_713804\|202 | ···················· | ···················· | ···················· | ···················· | ···················· | ···················· |
| X | 1577 | hCoV-19/Denmark/DCGC-15404/2020\|EPI_ISL_713805\|202 | ···················· | ···················· | ···················· | ···················· | ···················· | ···················· |
| X | 1578 | hCoV-19/Denmark/DCGC-15405/2020\|EPI_ISL_713806\|202 | ···················· | ···················· | ···················· | ···················· | ···················· | ···················· |
| X | 1579 | hCoV-19/Denmark/DCGC-15406/2020\|EPI_ISL_713807\|202 | ···················· | ·A·················· | ···················· | ···················· | ···················· | ··A················· |
| X | 1580 | hCoV-19/Denmark/DCGC-15408/2020\|EPI_ISL_713808\|202 | ···················· | ···················· | ···················· | ···················· | ···················· | ···················· |
| X | 1581 | hCoV-19/Denmark/DCGC-15409/2020\|EPI_ISL_713809\|202 | ···················· | ·A·················· | ···················· | ···················· | ···················· | ··A················· |
| X | 1582 | hCoV-19/Denmark/DCGC-15411/2020\|EPI_ISL_713810\|202 | ···················· | ···················· | ···················· | ···················· | ···················· | ···················· |
| X | 1583 | hCoV-19/Denmark/DCGC-15412/2020\|EPI_ISL_713811\|202 | ···················· | ···················· | ···················· | ···················· | ···················· | ···················· |
| X | 1584 | hCoV-19/Denmark/DCGC-15413/2020\|EPI_ISL_713812\|202 | ···················· | ···················· | ···················· | ···················· | ···················· | ···················· |
| X | 1585 | hCoV-19/Denmark/DCGC-15415/2020\|EPI_ISL_713813\|202 | ···················· | ···················· | ···················· | ···················· | ···················· | ···················· |
| X | 1586 | hCoV-19/Denmark/DCGC-15416/2020\|EPI_ISL_713814\|202 | ···················· | ···················· | ···················· | ···················· | ···················· | ···················· |
| X | 1587 | hCoV-19/Denmark/DCGC-15417/2020\|EPI_ISL_713815\|202 | ···················· | ·A·················· | ···················· | ···················· | ···················· | ···················· |
| X | 1588 | hCoV-19/Denmark/DCGC-15418/2020\|EPI_ISL_713816\|202 | ···················· | ···················· | ···················· | ···················· | ···················· | ···················· |
| X | 1589 | hCoV-19/Denmark/DCGC-15419/2020\|EPI_ISL_713817\|202 | ···················· | ···················· | ···················· | ···················· | ···················· | ···················· |
| X | 1590 | hCoV-19/Denmark/DCGC-15420/2020\|EPI_ISL_713818\|202 | ···················· | ···················· | ···················· | ···················· | ···················· | ···················· |
| X | 1591 | hCoV-19/Denmark/DCGC-15421/2020\|EPI_ISL_713819\|202 | ···················· | ···················· | ···················· | ···················· | ···················· | ···················· |
| X | 1592 | hCoV-19/Denmark/DCGC-15423/2020\|EPI_ISL_713820\|202 | ···················· | ···················· | ···················· | ···················· | ···················· | ···················· |
| X | 1593 | hCoV-19/Denmark/DCGC-15425/2020\|EPI_ISL_713821\|202 | ···················· | ···················· | ···················· | ···················· | ···················· | ···················· |
| X | 1594 | hCoV-19/Denmark/DCGC-15426/2020\|EPI_ISL_713822\|202 | ···················· | ···················· | ···················· | ···················· | ···················· | ···················· |
| X | 1595 | hCoV-19/Denmark/DCGC-15429/2020\|EPI_ISL_713823\|202 | ···················· | ···················· | ···················· | ···················· | ···················· | ···················· |
| X | 1596 | hCoV-19/Denmark/DCGC-15432/2020\|EPI_ISL_713824\|202 | ···················· | ···················· | ···················· | ···················· | ···················· | ···················· |
| X | 1597 | hCoV-19/Denmark/DCGC-15433/2020\|EPI_ISL_713825\|202 | ···················· | ···················· | ···················· | ···················· | ···················· | ···················· |
| X | 1598 | hCoV-19/Denmark/DCGC-15435/2020\|EPI_ISL_713826\|202 | ···················· | ···················· | ···················· | ···················· | ···················· | ···················· |
| X | 1599 | hCoV-19/Denmark/DCGC-15439/2020\|EPI_ISL_713827\|202 | ···················· | ·A·················· | ···················· | ···················· | ···················· | ··A················· |
| X | 1600 | hCoV-19/Denmark/DCGC-15440/2020\|EPI_ISL_713828\|202 | ···················· | ···················· | ···················· | ···················· | ···················· | ···················· |
| X | 1601 | hCoV-19/Denmark/DCGC-15441/2020\|EPI_ISL_713829\|202 | ···················· | ···················· | ···················· | ···················· | ···················· | ···················· |
| X | 1602 | hCoV-19/Denmark/DCGC-15449/2020\|EPI_ISL_713830\|202 | ···················· | ···················· | ···················· | ···················· | ···················· | ···················· |
| X | 1603 | hCoV-19/Denmark/DCGC-15451/2020\|EPI_ISL_713831\|202 | ···················· | ···················· | ···················· | ···················· | ···················· | ···················· |
| X | 1604 | hCoV-19/Denmark/DCGC-15452/2020\|EPI_ISL_713832\|202 | ···················· | ···················· | ···················· | ···················· | ···················· | ···················· |
| X | 1605 | hCoV-19/Denmark/DCGC-15455/2020\|EPI_ISL_713833\|202 | ···················· | ···················· | ···················· | ···················· | ···················· | ···················· |
| X | 1606 | hCoV-19/Denmark/DCGC-15458/2020\|EPI_ISL_713834\|202 | ···················· | ···················· | ···················· | ···················· | ···················· | ···················· |
| X | 1607 | hCoV-19/Denmark/DCGC-15460/2020\|EPI_ISL_713835\|202 | ···················· | ···················· | ···················· | ···················· | ···················· | ···················· |

| | | | | | | | | |
|---|---|---|---|---|---|---|---|---|
| X | 1608 | hCoV-19/Denmark/DCGC-15461/2020\|EPI_ISL_713836\|202 | ···················· | ···················· | ····················· | ···················· | ···················· | ···················· |
| X | 1609 | hCoV-19/Denmark/DCGC-15462/2020\|EPI_ISL_713837\|202 | ···················· | ···················· | ····················· | ···················· | ···················· | ···················· |
| X | 1610 | hCoV-19/Denmark/DCGC-15463/2020\|EPI_ISL_713838\|202 | ···················· | ···················· | ····················· | ···················· | ···················· | ···················· |
| X | 1611 | hCoV-19/Denmark/DCGC-15464/2020\|EPI_ISL_713839\|202 | ···················· | ···················· | ····················· | ···················· | ···················· | ···················· |
| X | 1612 | hCoV-19/Denmark/DCGC-15467/2020\|EPI_ISL_713840\|202 | ···················· | ···················· | ····················· | ···················· | ···················· | ···················· |
| X | 1613 | hCoV-19/Denmark/DCGC-15469/2020\|EPI_ISL_713841\|202 | ···················· | ···················· | ····················· | ···················· | ···················· | ···················· |
| X | 1614 | hCoV-19/Denmark/DCGC-15471/2020\|EPI_ISL_713842\|202 | ···················· | ···················· | ····················· | ···················· | ···················· | ···················· |
| X | 1615 | hCoV-19/Denmark/DCGC-15473/2020\|EPI_ISL_713843\|202 | ···················· | ···················· | ····················· | ···················· | ···················· | ···················· |
| X | 1616 | hCoV-19/Denmark/DCGC-15475/2020\|EPI_ISL_713844\|202 | ···················· | ···················· | ····················· | ···················· | ···················· | ···················· |
| X | 1617 | hCoV-19/Denmark/DCGC-15476/2020\|EPI_ISL_713845\|202 | ···················· | ···················· | ····················· | ···················· | ···················· | ···················· |
| X | 1618 | hCoV-19/Denmark/DCGC-15479/2020\|EPI_ISL_713846\|202 | ···················· | ···················· | ····················· | ···················· | ···················· | ···················· |
| X | 1619 | hCoV-19/Denmark/DCGC-15481/2020\|EPI_ISL_713847\|202 | ···················· | ···················· | ····················· | ···················· | ···················· | ···················· |
| X | 1620 | hCoV-19/Denmark/DCGC-15483/2020\|EPI_ISL_713848\|202 | ···················· | ···················· | ····················· | ···················· | ···················· | ···················· |
| X | 1621 | hCoV-19/Denmark/DCGC-15484/2020\|EPI_ISL_713849\|202 | ···················· | ···················· | ····················· | ···················· | ···················· | ···················· |
| X | 1622 | hCoV-19/Denmark/DCGC-15486/2020\|EPI_ISL_713850\|202 | ···················· | ···················· | ····················· | ···················· | ···················· | ···················· |
| X | 1623 | hCoV-19/Denmark/DCGC-15489/2020\|EPI_ISL_713851\|202 | ···················· | ···················· | ····················· | ···················· | ···················· | ···················· |
| X | 1624 | hCoV-19/Denmark/DCGC-15491/2020\|EPI_ISL_713852\|202 | ···················· | ···················· | ····················· | ···················· | ···················· | ···················· |
| X | 1625 | hCoV-19/Denmark/DCGC-15495/2020\|EPI_ISL_713853\|202 | ···················· | ···················· | ····················· | ···················· | ···················· | ···················· |
| X | 1626 | hCoV-19/Denmark/DCGC-15496/2020\|EPI_ISL_713854\|202 | ···················· | ···················· | ····················· | ···················· | ······T············· | ···················· |
| X | 1627 | hCoV-19/Denmark/DCGC-15503/2020\|EPI_ISL_713855\|202 | ···················· | ···················· | ····················· | ···················· | ···················· | ···················· |
| X | 1628 | hCoV-19/Denmark/DCGC-15504/2020\|EPI_ISL_713856\|202 | ···················· | ···················· | ····················· | ···················· | ···················· | ···················· |
| X | 1629 | hCoV-19/Denmark/DCGC-15506/2020\|EPI_ISL_713857\|202 | ···················· | ···················· | ····················· | ···················· | ···················· | ···················· |
| X | 1630 | hCoV-19/Denmark/DCGC-15547/2020\|EPI_ISL_713883\|202 | ···················· | ···················· | ····················· | ···················· | ···················· | ···················· |
| X | 1631 | hCoV-19/Denmark/DCGC-15549/2020\|EPI_ISL_713884\|202 | ···················· | ···················· | ····················· | ···················· | ······T············· | ···················· |
| X | 1632 | hCoV-19/Denmark/DCGC-15550/2020\|EPI_ISL_713885\|202 | ···················· | ···················· | ····················· | ···················· | ···················· | ···················· |
| X | 1633 | hCoV-19/Denmark/DCGC-15551/2020\|EPI_ISL_713886\|202 | ···················· | ···················· | ····················· | ···················· | ···················· | ···················· |
| X | 1634 | hCoV-19/Denmark/DCGC-15552/2020\|EPI_ISL_713887\|202 | ···················· | ···················· | ····················· | ···················· | ···················· | ···················· |
| X | 1635 | hCoV-19/Denmark/DCGC-15554/2020\|EPI_ISL_713888\|202 | ···················· | ···················· | ····················· | ···················· | ···················· | ···················· |
| X | 1636 | hCoV-19/Denmark/DCGC-15556/2020\|EPI_ISL_713889\|202 | ···················· | ···················· | ····················· | ···················· | ···················· | ···················· |
| X | 1637 | hCoV-19/England/CAMC-C44DEF/2020\|EPI_ISL_718600\|20 | ···················· | ···················· | ····················· | ···················· | ···················· | ···················· |
| X | 1638 | hCoV-19/England/CAMC-C46AB9/2020\|EPI_ISL_718605\|20 | ···················· | ···················· | ····················· | ···················· | ···················· | ···················· |
| X | 1639 | hCoV-19/England/CAMC-C467EF/2020\|EPI_ISL_718607\|20 | ···················· | ·A·················· | ····················· | ···················· | ···················· | ···················· |
| X | 1640 | hCoV-19/England/CAMC-C45A8D/2020\|EPI_ISL_718608\|20 | ···················· | ···················· | ····················· | ···················· | ···················· | ···················· |
| X | 1641 | hCoV-19/England/CAMC-C44C88/2020\|EPI_ISL_718609\|20 | ···················· | ···················· | ····················· | ···················· | ···················· | ···················· |
| X | 1642 | hCoV-19/England/CAMC-C4762D/2020\|EPI_ISL_718630\|20 | ···················· | ···················· | ····················· | ···················· | ···················· | ···················· |
| X | 1643 | hCoV-19/England/ALDP-C49CDE/2020\|EPI_ISL_718631\|20 | ···················· | ···················· | ····················· | ···················· | ···················· | ···················· |
| X | 1644 | hCoV-19/England/ALDP-C480B5/2020\|EPI_ISL_718637\|20 | ···················· | ···················· | ····················· | ···················· | ···················· | ···················· |
| X | 1645 | hCoV-19/England/CAMC-C46BC5/2020\|EPI_ISL_718638\|20 | ···················· | ···················· | ····················· | ···················· | ···················· | ···················· |
| X | 1646 | hCoV-19/England/ALDP-C49BD1/2020\|EPI_ISL_718639\|20 | ···················· | ···················· | ····················· | ···················· | ···················· | ···················· |
| X | 1647 | hCoV-19/England/ALDP-C49C92/2020\|EPI_ISL_718640\|20 | ···················· | ···················· | ····················· | ···················· | ······T············· | ···················· |
| X | 1648 | hCoV-19/England/ALDP-C49B0E/2020\|EPI_ISL_718643\|20 | ···················· | ···················· | ····················· | ···················· | ···················· | ···················· |
| X | 1649 | hCoV-19/England/ALDP-C47EBC/2020\|EPI_ISL_718644\|20 | ···················· | ···················· | ····················· | ···················· | ···················· | ···················· |
| X | 1650 | hCoV-19/England/ALDP-C47F04/2020\|EPI_ISL_718647\|20 | ···················· | ···················· | ····················· | ···················· | ···················· | ···················· |
| X | 1651 | hCoV-19/England/CAMC-C47010/2020\|EPI_ISL_718648\|20 | ···················· | ···················· | ····················· | ···················· | ···················· | ···················· |
| X | 1652 | hCoV-19/England/ALDP-C49C65/2020\|EPI_ISL_718650\|20 | ···················· | ···················· | ····················· | ···················· | ···················· | ···················· |
| X | 1653 | hCoV-19/England/ALDP-C498CB/2020\|EPI_ISL_718653\|20 | ···················· | ···················· | ····················· | ···················· | ···················· | ···················· |
| X | 1654 | hCoV-19/England/CAMC-C4713B/2020\|EPI_ISL_718655\|20 | ···················· | ···················· | ····················· | ···················· | ···················· | ···················· |
| X | 1655 | hCoV-19/England/ALDP-C48158/2020\|EPI_ISL_718657\|20 | ···················· | ···················· | ····················· | ···················· | ···················· | ···················· |
| X | 1656 | hCoV-19/England/ALDP-C47C85/2020\|EPI_ISL_718658\|20 | ···················· | ···················· | ····················· | ···················· | ···················· | ···················· |
| X | 1657 | hCoV-19/England/ALDP-C498BC/2020\|EPI_ISL_718662\|20 | ···················· | ···················· | ····················· | ···················· | ···················· | ···················· |
| X | 1658 | hCoV-19/England/ALDP-C47D0A/2020\|EPI_ISL_718663\|20 | ···················· | ···················· | ····················· | ···················· | ···················· | ···················· |
| X | 1659 | hCoV-19/England/ALDP-C47D37/2020\|EPI_ISL_718664\|20 | ···················· | ···················· | ····················· | ···················· | ···················· | ···················· |
| X | 1660 | hCoV-19/England/ALDP-C49807/2020\|EPI_ISL_718666\|20 | ···················· | ···················· | ····················· | ···················· | ···················· | ···A················ |
| X | 1661 | hCoV-19/England/CAMC-C47511/2020\|EPI_ISL_718667\|20 | ···················· | ···················· | ····················· | ···················· | ···················· | ···················· |
| X | 1662 | hCoV-19/England/CAMC-C47854/2020\|EPI_ISL_718668\|20 | ···················· | ···················· | ····················· | ···················· | ···················· | ···················· |
| X | 1663 | hCoV-19/England/ALDP-C49A89/2020\|EPI_ISL_718670\|20 | ···················· | ···················· | ····················· | ···················· | ···················· | ···················· |
| X | 1664 | hCoV-19/England/ALDP-C47EDA/2020\|EPI_ISL_718672\|20 | ···················· | ···················· | ····················· | ···················· | ···················· | ···················· |
| X | 1665 | hCoV-19/England/ALDP-C4811C/2020\|EPI_ISL_718674\|20 | ···················· | ···················· | ····················· | ···················· | ···················· | ···················· |
| X | 1666 | hCoV-19/England/ALDP-C49861/2020\|EPI_ISL_718676\|20 | ···················· | ···················· | ····················· | ···················· | ···················· | ···················· |
| X | 1667 | hCoV-19/England/ALDP-C48167/2020\|EPI_ISL_718678\|20 | ···················· | ···················· | ····················· | ···················· | ···················· | ···················· |
| X | 1668 | hCoV-19/England/CAMC-C47283/2020\|EPI_ISL_718679\|20 | ···················· | ···················· | ····················· | ···················· | ······T············· | ···················· |
| X | 1669 | hCoV-19/England/CAMC-C46C59/2020\|EPI_ISL_718681\|20 | ···················· | ····A··············· | ····················· | ···················· | ···················· | ···················· |
| X | 1670 | hCoV-19/England/CAMC-C4755D/2020\|EPI_ISL_718683\|20 | ···················· | ···················· | ····················· | ···················· | ···················· | ···················· |
| X | 1671 | hCoV-19/England/CAMC-C472A1/2020\|EPI_ISL_718684\|20 | ···················· | ···················· | ····················· | ···················· | ···················· | ···················· |
| X | 1672 | hCoV-19/England/CAMC-C473EA/2020\|EPI_ISL_718685\|20 | ···················· | ···················· | ····················· | ···················· | ···················· | ···················· |
| X | 1673 | hCoV-19/England/ALDP-C47EE9/2020\|EPI_ISL_718686\|20 | ···················· | ···················· | ····················· | ···················· | ···················· | ···················· |
| X | 1674 | hCoV-19/England/ALDP-C4805B/2020\|EPI_ISL_718687\|20 | ···················· | ···················· | ····················· | ···················· | ···················· | ···················· |
| X | 1675 | hCoV-19/England/CAMC-C477FD/2020\|EPI_ISL_718688\|20 | ···················· | ···················· | ····················· | ···················· | ···················· | ···················· |
| X | 1676 | hCoV-19/England/ALDP-C47F8C/2020\|EPI_ISL_718691\|20 | ···················· | ···················· | ····················· | ············A······· | ···················· | ···················· |
| X | 1677 | hCoV-19/England/QEUH-BA1BC9/2020\|EPI_ISL_719922\|20 | ···················· | ···················· | ····················· | ···················· | ···················· | ···················· |
| X | 1678 | hCoV-19/England/QEUH-B9EE02/2020\|EPI_ISL_719923\|20 | ···················· | ···················· | ····················· | ···················· | ···················· | ···················· |
| X | 1679 | hCoV-19/England/QEUH-BA27F1/2020\|EPI_ISL_719924\|20 | ···················· | ···················· | ····················· | ···················· | ···················· | ···················· |
| X | 1680 | hCoV-19/England/QEUH-B9E6EC/2020\|EPI_ISL_719925\|20 | ···················· | ···················· | ····················· | ···················· | ···················· | ···················· |
| X | 1681 | hCoV-19/England/QEUH-B9E6DD/2020\|EPI_ISL_719927\|20 | ···················· | ···················· | ····················· | ···················· | ···················· | ···················· |
| X | 1682 | hCoV-19/England/QEUH-BA1C4E/2020\|EPI_ISL_719928\|20 | ···················· | ···················· | ····················· | ···················· | ······T············· | ···················· |
| X | 1683 | hCoV-19/England/QEUH-B9F01A/2020\|EPI_ISL_719929\|20 | ···················· | ···················· | ····················· | ···················· | ···················· | ···················· |
| X | 1684 | hCoV-19/England/QEUH-B9F0CF/2020\|EPI_ISL_719930\|20 | ···················· | ···················· | ····················· | ···················· | ···················· | ···················· |
| X | 1685 | hCoV-19/England/QEUH-B9F0FC/2020\|EPI_ISL_719931\|20 | ···················· | ···················· | ····················· | ···················· | ···················· | ···················· |
| X | 1686 | hCoV-19/England/QEUH-B9ED9C/2020\|EPI_ISL_719934\|20 | ···················· | ···················· | ····················· | ···················· | ···················· | ···················· |
| X | 1687 | hCoV-19/England/QEUH-B9E804/2020\|EPI_ISL_719938\|20 | ···················· | ···················· | ····················· | ···················· | ···················· | ···················· |

| | | | | | | | | |
|---|---|---|---|---|---|---|---|---|
| X | 1688 | hCoV-19/Wales/QEUH-BA1C5D/2020\|EPI_ISL_719961\|2020 | ···················· | ···················· | ····················· | ···················· | ···················· | ···················· |
| X | 1689 | hCoV-19/England/QEUH-BA2199/2020\|EPI_ISL_719963\|20 | ···················· | ···················· | ····················· | ···················· | ···················· | ···················· |
| X | 1690 | hCoV-19/England/QEUH-B9E6CE/2020\|EPI_ISL_719964\|20 | ···················· | ·A·················· | ····················· | ···················· | ···················· | ···A················ |
| X | 1691 | hCoV-19/England/QEUH-BA273D/2020\|EPI_ISL_719965\|20 | ···················· | ···················· | ····················· | ···················· | ···················· | ···················· |
| X | 1692 | hCoV-19/England/QEUH-BA1C7B/2020\|EPI_ISL_719966\|20 | ···················· | ···················· | ····················· | ···················· | ···················· | ···················· |
| X | 1693 | hCoV-19/England/QEUH-BA208D/2020\|EPI_ISL_719969\|20 | ···················· | ···················· | ····················· | ···················· | ···················· | ···················· |
| X | 1694 | hCoV-19/England/QEUH-B9F19F/2020\|EPI_ISL_719972\|20 | ···················· | ···················· | ····················· | ···················· | ···················· | ···················· |
| X | 1695 | hCoV-19/England/QEUH-BA1C11/2020\|EPI_ISL_719974\|20 | ···················· | ···················· | ····················· | ···················· | ···················· | ···A················ |
| X | 1696 | hCoV-19/England/QEUH-BA23A2/2020\|EPI_ISL_719975\|20 | ···················· | ···················· | ····················· | ···················· | ···················· | ···················· |
| X | 1697 | hCoV-19/England/QEUH-BA1F63/2020\|EPI_ISL_719976\|20 | ········Y··········· | ···················· | ····················· | ···················· | ···················· | ···················· |
| X | 1698 | hCoV-19/England/QEUH-BA26F4/2020\|EPI_ISL_719978\|20 | ···················· | ···················· | ····················· | ···················· | ···················· | ···················· |
| X | 1699 | hCoV-19/England/QEUH-B9EDD8/2020\|EPI_ISL_719979\|20 | ···················· | ···················· | ····················· | ···················· | ···················· | ···················· |
| X | 1700 | hCoV-19/England/QEUH-BA22A5/2020\|EPI_ISL_719980\|20 | ···················· | ···················· | ····················· | ···················· | ···················· | ···················· |
| X | 1701 | hCoV-19/England/QEUH-BA1FAF/2020\|EPI_ISL_719981\|20 | ···K················ | ···················· | ····················· | ···················· | ···················· | ···················· |
| X | 1702 | hCoV-19/England/QEUH-B9E707/2020\|EPI_ISL_719983\|20 | ···················· | ···················· | ····················· | ···················· | ···················· | ···················· |
| X | 1703 | hCoV-19/England/QEUH-B9F0B0/2020\|EPI_ISL_719984\|20 | ···················· | ···················· | ····················· | ···················· | ···················· | ···················· |
| X | 1704 | hCoV-19/England/QEUH-B9E725/2020\|EPI_ISL_719985\|20 | ···················· | ···················· | ····················· | ···················· | ···················· | ···················· |
| X | 1705 | hCoV-19/England/QEUH-B9F092/2020\|EPI_ISL_719986\|20 | ···················· | ···················· | ····················· | ···················· | ···················· | ···················· |
| X | 1706 | hCoV-19/England/MILK-BB1908/2020\|EPI_ISL_719991\|20 | ···················· | ···················· | ····················· | ···················· | ···················· | ···················· |
| X | 1707 | hCoV-19/England/MILK-BB1258/2020\|EPI_ISL_719995\|20 | ···················· | ···················· | ····················· | ···················· | ···················· | ···················· |
| X | 1708 | hCoV-19/England/MILK-BB1E27/2020\|EPI_ISL_719996\|20 | ···················· | ···················· | ····················· | ···················· | ···················· | ···················· |
| X | 1709 | hCoV-19/England/MILK-BB1267/2020\|EPI_ISL_719997\|20 | ···················· | ···················· | ····················· | ···················· | ···················· | ···················· |
| X | 1710 | hCoV-19/England/MILK-BB0FCB/2020\|EPI_ISL_719998\|20 | ···················· | ···················· | ····················· | ···················· | ···················· | ···················· |
| X | 1711 | hCoV-19/England/MILK-BFCE02/2020\|EPI_ISL_720902\|20 | ···················· | ···················· | ····················· | ···················· | ···················· | ···················· |
| X | 1712 | hCoV-19/England/QEUH-C01443/2020\|EPI_ISL_720904\|20 | ···················· | ···················· | ····················· | ···················· | ···················· | ···················· |
| X | 1713 | hCoV-19/England/MILK-BFCB65/2020\|EPI_ISL_720906\|20 | ···················· | ···················· | ····················· | ···················· | ···················· | ···················· |
| X | 1714 | hCoV-19/Wales/ALDP-C048EA/2020\|EPI_ISL_720907\|2020 | ···················· | ···················· | ····················· | ···················· | ···················· | ···················· |
| X | 1715 | hCoV-19/England/MILK-BFCF4B/2020\|EPI_ISL_720908\|20 | ···················· | ···················· | ····················· | ···················· | ···················· | ···················· |
| X | 1716 | hCoV-19/England/QEUH-B9F96A/2020\|EPI_ISL_720909\|20 | ···················· | ··R················· | ····················· | ···················· | ···················· | ···················· |
| X | 1717 | hCoV-19/England/QEUH-B9F44B/2020\|EPI_ISL_720910\|20 | ···················· | ···················· | ····················· | ···················· | ···················· | ···················· |
| X | 1718 | hCoV-19/England/QEUH-B9F618/2020\|EPI_ISL_720911\|20 | ···················· | ···················· | ····················· | ···················· | ···················· | ···················· |
| X | 1719 | hCoV-19/England/QEUH-BA3259/2020\|EPI_ISL_720912\|20 | ···················· | ···················· | ····················· | ···················· | ···················· | ···················· |
| X | 1720 | hCoV-19/England/QEUH-B9F6FA/2020\|EPI_ISL_720913\|20 | ···················· | ···················· | ····················· | ···················· | ···················· | ···················· |
| X | 1721 | hCoV-19/England/QEUH-B9F94C/2020\|EPI_ISL_720914\|20 | ···················· | ···················· | ····················· | ···················· | ···················· | ···················· |
| X | 1722 | hCoV-19/England/QEUH-B9E42E/2020\|EPI_ISL_720915\|20 | ···················· | ···················· | ····················· | ···················· | ···················· | ···················· |
| X | 1723 | hCoV-19/England/QEUH-B9FBA0/2020\|EPI_ISL_720917\|20 | ···················· | ·A·················· | ····················· | ···················· | ···················· | ···A················ |
| X | 1724 | hCoV-19/England/QEUH-B9FC07/2020\|EPI_ISL_720918\|20 | ···················· | ·A·················· | ····················· | ···················· | ···················· | ···A················ |
| X | 1725 | hCoV-19/England/QEUH-B9F6EB/2020\|EPI_ISL_720919\|20 | ···················· | ···················· | ····················· | ···················· | ···················· | ···················· |
| X | 1726 | hCoV-19/England/QEUH-B9F7D9/2020\|EPI_ISL_720942\|20 | ···················· | ···················· | ····················· | ···················· | ···················· | ···················· |
| X | 1727 | hCoV-19/England/QEUH-B9F609/2020\|EPI_ISL_720943\|20 | ···················· | ···················· | ····················· | ···················· | ···················· | ···················· |
| X | 1728 | hCoV-19/England/QEUH-B9FE3E/2020\|EPI_ISL_720944\|20 | ···················· | ···················· | ····················· | ···················· | ···················· | ···················· |
| X | 1729 | hCoV-19/England/QEUH-B9FE01/2020\|EPI_ISL_720945\|20 | ···················· | ···················· | ····················· | ···················· | ···················· | ···················· |
| X | 1730 | hCoV-19/England/QEUH-BA3D1C/2020\|EPI_ISL_720946\|20 | ···················· | ···················· | ····················· | ···················· | ···················· | ···················· |
| X | 1731 | hCoV-19/England/QEUH-B9F627/2020\|EPI_ISL_720948\|20 | ···················· | ···················· | ····················· | ···················· | ···················· | ···················· |
| X | 1732 | hCoV-19/England/QEUH-B9FCBC/2020\|EPI_ISL_720949\|20 | ···················· | ···················· | ····················· | ···················· | ···················· | ···················· |
| X | 1733 | hCoV-19/England/QEUH-B9E0C0/2020\|EPI_ISL_720950\|20 | ···················· | ···················· | ····················· | ···················· | ···················· | ···················· |
| X | 1734 | hCoV-19/England/QEUH-B9E206/2020\|EPI_ISL_720952\|20 | ···················· | ···················· | ····················· | ···················· | ···················· | ···················· |
| X | 1735 | hCoV-19/England/QEUH-B9F77F/2020\|EPI_ISL_720953\|20 | ···················· | ···················· | ····················· | ···················· | ···················· | ···················· |
| X | 1736 | hCoV-19/England/QEUH-B9F95B/2020\|EPI_ISL_720955\|20 | ···················· | ···················· | ····················· | ···················· | ···················· | ···················· |
| X | 1737 | hCoV-19/England/QEUH-BA2CF2/2020\|EPI_ISL_720957\|20 | ···················· | ···················· | ····················· | ···················· | ···················· | ···················· |
| X | 1738 | hCoV-19/England/QEUH-B9F706/2020\|EPI_ISL_720959\|20 | ···················· | ···················· | ····················· | ···················· | ···················· | ···················· |
| X | 1739 | hCoV-19/England/QEUH-B9F8C7/2020\|EPI_ISL_720960\|20 | ···················· | ···················· | ····················· | ···················· | ···················· | ···················· |
| X | 1740 | hCoV-19/England/QEUH-BA3909/2020\|EPI_ISL_720961\|20 | ···················· | ···················· | ····················· | ···················· | ···················· | ···················· |
| X | 1741 | hCoV-19/England/QEUH-B9E37C/2020\|EPI_ISL_720962\|20 | ···················· | ···················· | ····················· | ···················· | ···················· | ···················· |
| X | 1742 | hCoV-19/England/QEUH-BA3480/2020\|EPI_ISL_720963\|20 | ···················· | ···················· | ····················· | ···················· | ···················· | ···················· |
| X | 1743 | hCoV-19/England/QEUH-B9FB64/2020\|EPI_ISL_720964\|20 | ···················· | ···················· | ····················· | ···················· | ···················· | ···················· |
| X | 1744 | hCoV-19/England/QEUH-B9F4F0/2020\|EPI_ISL_720965\|20 | ···················· | ···················· | ····················· | ···················· | ···················· | ···················· |
| X | 1745 | hCoV-19/England/QEUH-B9F584/2020\|EPI_ISL_720966\|20 | ···················· | ···················· | ····················· | ···················· | ···················· | ···················· |
| X | 1746 | hCoV-19/England/QEUH-B9FB28/2020\|EPI_ISL_720967\|20 | ···················· | ···················· | ····················· | ···················· | ···················· | ···················· |
| X | 1747 | hCoV-19/England/QEUH-B9E39A/2020\|EPI_ISL_720969\|20 | ···················· | ···················· | ····················· | ···················· | ···················· | ···················· |
| X | 1748 | hCoV-19/England/QEUH-BA2D95/2020\|EPI_ISL_720970\|20 | ···················· | ······A············· | ····················· | ···················· | ···················· | ···················· |
| X | 1749 | hCoV-19/England/QEUH-B9F830/2020\|EPI_ISL_720971\|20 | ···················· | ···················· | ····················· | ···················· | ···················· | ···················· |
| X | 1750 | hCoV-19/England/QEUH-B9F399/2020\|EPI_ISL_720972\|20 | ···················· | ···················· | ····················· | ···················· | ···················· | ···················· |
| X | 1751 | hCoV-19/England/QEUH-B9FA76/2020\|EPI_ISL_720974\|20 | ···················· | ···················· | ····················· | ···················· | ···················· | ···················· |
| X | 1752 | hCoV-19/England/QEUH-B9FB91/2020\|EPI_ISL_720975\|20 | ···················· | ···················· | ····················· | ···················· | ···················· | ···················· |
| X | 1753 | hCoV-19/England/QEUH-B9F50C/2020\|EPI_ISL_720976\|20 | ···················· | ···················· | ····················· | ···················· | ···················· | ···················· |
| X | 1754 | hCoV-19/England/QEUH-B9FA58/2020\|EPI_ISL_720977\|20 | ···················· | ···················· | ····················· | ···················· | ···················· | ···················· |
| X | 1755 | hCoV-19/England/QEUH-B9F29C/2020\|EPI_ISL_720978\|20 | ···················· | ···················· | ····················· | ···················· | ···················· | ···················· |
| X | 1756 | hCoV-19/England/QEUH-BA41E2/2020\|EPI_ISL_720979\|20 | ···················· | ···················· | ····················· | ···················· | ···················· | ···················· |
| X | 1757 | hCoV-19/England/QEUH-B9E34F/2020\|EPI_ISL_720980\|20 | ···················· | ·A·················· | ····················· | ···················· | ··········T········· | ···················· |
| X | 1758 | hCoV-19/England/QEUH-BA3D3A/2020\|EPI_ISL_720981\|20 | ···················· | ···················· | ····················· | ···················· | ······T············· | ···················· |
| X | 1759 | hCoV-19/England/QEUH-B9F672/2020\|EPI_ISL_720982\|20 | ···················· | ···················· | ····················· | ···················· | ···················· | ···················· |
| X | 1760 | hCoV-19/England/QEUH-B9E136/2020\|EPI_ISL_720983\|20 | ···················· | ···················· | ····················· | ···················· | ···················· | ···················· |
| X | 1761 | hCoV-19/England/QEUH-BA422B/2020\|EPI_ISL_720984\|20 | ···················· | ···················· | ····················· | ···················· | ···················· | ···················· |
| X | 1762 | hCoV-19/England/QEUH-B9F88B/2020\|EPI_ISL_720985\|20 | ···················· | ···················· | ····················· | ···················· | ···················· | ···················· |
| X | 1763 | hCoV-19/England/QEUH-B9E00C/2020\|EPI_ISL_720986\|20 | ···················· | ···················· | ····················· | ···················· | ···················· | ···················· |
| X | 1764 | hCoV-19/England/QEUH-BA39AF/2020\|EPI_ISL_720988\|20 | ···················· | ···················· | ····················· | ···················· | ···················· | ···················· |
| X | 1765 | hCoV-19/England/QEUH-B9E109/2020\|EPI_ISL_720990\|20 | ···················· | ·A·················· | ····················· | ···················· | ···················· | ···················· |
| X | 1766 | hCoV-19/England/QEUH-B9FB0A/2020\|EPI_ISL_720991\|20 | ···················· | ···················· | ····················· | ···················· | ···················· | ···················· |
| X | 1767 | hCoV-19/England/QEUH-BA34EA/2020\|EPI_ISL_720993\|20 | ···················· | ···················· | ····················· | ···················· | ···················· | ···················· |

X 1768 hCoV-19/England/QEUH-B9F34E/2020|EPI_ISL_720994|20 ···················· ···················· ····················· ···················· ···················· ····················
X 1769 hCoV-19/England/QEUH-B9FB19/2020|EPI_ISL_720995|20 ···················· ···················· ····················· ···················· ···················· ····················
X 1770 hCoV-19/England/QEUH-BA35AB/2020|EPI_ISL_720996|20 ···················· ···················· ····················· ···················· ···················· ····················
X 1771 hCoV-19/England/QEUH-B9F2E7/2020|EPI_ISL_720998|20 ···················· ···················· ····················· ···················· ···················· ···············A····
X 1772 hCoV-19/Netherlands/OV-RIVM-20314/2020|EPI_ISL_723 ···················· ···················· ····················· ···················· ···················· ····················
X 1773 hCoV-19/Netherlands/GE-RIVM-20317/2020|EPI_ISL_723 ···················· ···················· ····················· ···················· ···················· ····················
X 1774 hCoV-19/Netherlands/GE-RIVM-20320/2020|EPI_ISL_723 ···················· ···················· ····················· ···················· ···················· ····················
X 1775 hCoV-19/Netherlands/GE-RIVM-20324/2020|EPI_ISL_723 ···················· ···················· ····················· ···················· ···················· ····················
X 1776 hCoV-19/Netherlands/GE-RIVM-20330/2020|EPI_ISL_723 ···················· ···················· ····················· ···················· ···················· ····················
X 1777 hCoV-19/Netherlands/GE-RIVM-20331/2020|EPI_ISL_723 ···················· ···················· ····················· ···················· ···················· ····················
X 1778 hCoV-19/Netherlands/GE-RIVM-20333/2020|EPI_ISL_723 ···················· ···················· ····················· ···················· ···················· ····················
X 1779 hCoV-19/Netherlands/GE-RIVM-20334/2020|EPI_ISL_723 ···················· ···················· ····················· ···················· ···················· ····················
X 1780 hCoV-19/Netherlands/GE-RIVM-20336/2020|EPI_ISL_723 ···················· ···················· ····················· ···················· ···················· ····················
X 1781 hCoV-19/Netherlands/GE-RIVM-20337/2020|EPI_ISL_723 ···················· ···················· ····················· ···················· ···················· ····················
X 1782 hCoV-19/Netherlands/GE-RIVM-20339/2020|EPI_ISL_723 ···················· ···················· ····················· ···················· ···················· ····················
X 1783 hCoV-19/Netherlands/GE-RIVM-20341/2020|EPI_ISL_723 ···················· ···················· ····················· ···················· ···················· ····················
X 1784 hCoV-19/Netherlands/UT-RIVM-20132/2020|EPI_ISL_723 ···················· ···················· ····················· ···················· ···················· ····················
X 1785 hCoV-19/Netherlands/NH-RIVM-20221/2020|EPI_ISL_723 ···················· ·A·················· ····················· ···················· ···················· ···A················
X 1786 hCoV-19/Netherlands/NH-RIVM-20343/2020|EPI_ISL_723 ···················· ···················· ····················· ···················· ···················· ····················
X 1787 hCoV-19/Netherlands/NH-RIVM-20003/2020|EPI_ISL_723 ···················· ···················· ····················· ···················· ···················· ····················
X 1788 hCoV-19/Netherlands/NH-RIVM-20004/2020|EPI_ISL_723 ···················· ···················· ····················· ···················· ···················· ····················
X 1789 hCoV-19/Netherlands/NH-RIVM-20005/2020|EPI_ISL_723 ···················· ···················· ····················· ···················· ···················· ····················
X 1790 hCoV-19/Netherlands/NH-RIVM-20006/2020|EPI_ISL_723 ···················· ···················· ····················· ···················· ···················· ····················
X 1791 hCoV-19/Netherlands/NH-RIVM-20007/2020|EPI_ISL_723 ···················· ···················· ····················· ···················· ······T············· ····················
X 1792 hCoV-19/Netherlands/NH-RIVM-20008/2020|EPI_ISL_723 ···················· ···················· ····················· ···················· ······T············· ····················
X 1793 hCoV-19/Denmark/DCGC-13982/2020|EPI_ISL_711233|202 ···················· ···················· ····················· ···················· ···················· ····················
X 1794 hCoV-19/Denmark/DCGC-14039/2020|EPI_ISL_711234|202 ···················· ···················· ····················· ···················· ···················· ····················
X 1795 hCoV-19/Denmark/DCGC-14048/2020|EPI_ISL_711235|202 ···················· ···················· ····················· ···················· ···················· ····················
X 1796 hCoV-19/Denmark/DCGC-14056/2020|EPI_ISL_711236|202 ···················· ···················· ····················· ···················· ···················· ····················
X 1797 hCoV-19/Denmark/DCGC-14106/2020|EPI_ISL_711237|202 ···················· ···················· ····················· ···················· ···················· ····················
X 1798 hCoV-19/Denmark/DCGC-14112/2020|EPI_ISL_711238|202 ···················· ···················· ····················· ···················· ···················· ····················
X 1799 hCoV-19/Denmark/DCGC-14149/2020|EPI_ISL_711239|202 ···················· ···················· ····················· ···················· ···················· ····················
X 1800 hCoV-19/Denmark/DCGC-14167/2020|EPI_ISL_711240|202 ···················· ···················· ····················· ···················· ···················· ····················
X 1801 hCoV-19/Denmark/DCGC-14168/2020|EPI_ISL_711241|202 ···················· ···················· ····················· ···················· ···················· ····················
X 1802 hCoV-19/Denmark/DCGC-14293/2020|EPI_ISL_711242|202 ···················· ···················· ····················· ···················· ···················· ····················
X 1803 hCoV-19/Denmark/DCGC-14303/2020|EPI_ISL_711243|202 ···················· ···················· ····················· ···················· ···················· ····················
X 1804 hCoV-19/Denmark/DCGC-14318/2020|EPI_ISL_711244|202 ···················· ···················· ····················· ···················· ···················· ····················
X 1805 hCoV-19/Denmark/DCGC-14333/2020|EPI_ISL_711245|202 ···················· ···················· ····················· ···················· ···················· ····················
X 1806 hCoV-19/Denmark/DCGC-14390/2020|EPI_ISL_711246|202 ···················· ···················· ····················· ···················· ···················· ····················
X 1807 hCoV-19/Denmark/DCGC-14393/2020|EPI_ISL_711247|202 ···················· ···················· ····················· ···················· ···················· ····················
X 1808 hCoV-19/Denmark/DCGC-14403/2020|EPI_ISL_711248|202 ···················· ···················· ····················· ···················· ···················· ····················
X 1809 hCoV-19/Denmark/DCGC-14432/2020|EPI_ISL_711249|202 ···················· ···················· ····················· ···················· ···················· ····················
X 1810 hCoV-19/Denmark/DCGC-14446/2020|EPI_ISL_711250|202 ···················· ···················· ····················· ···················· ···················· ····················
X 1811 hCoV-19/Denmark/DCGC-14451/2020|EPI_ISL_711251|202 ···················· ···················· ····················· ···················· ···················· ····················
X 1812 hCoV-19/Denmark/DCGC-14479/2020|EPI_ISL_711252|202 ···················· ···················· ····················· ···················· ···················· ····················
X 1813 hCoV-19/Denmark/DCGC-14489/2020|EPI_ISL_711253|202 ···················· ···················· ····················· ···················· ···················· ····················
X 1814 hCoV-19/Denmark/DCGC-14497/2020|EPI_ISL_711254|202 ···················· ···················· ····················· ···················· ···················· ····················
X 1815 hCoV-19/Denmark/DCGC-14529/2020|EPI_ISL_711255|202 ···················· ···················· ····················· ···················· ···················· ····················
X 1816 hCoV-19/Denmark/DCGC-14594/2020|EPI_ISL_711256|202 ···················· ···················· ····················· ···················· ···················· ····················
X 1817 hCoV-19/Denmark/DCGC-14616/2020|EPI_ISL_711257|202 ···················· ···················· ····················· ···················· ···················· ····················
X 1818 hCoV-19/Denmark/DCGC-14619/2020|EPI_ISL_711258|202 ···················· ···················· ····················· ···················· ···················· ····················
X 1819 hCoV-19/Denmark/DCGC-14646/2020|EPI_ISL_711259|202 ···················· ···················· ····················· ···················· ···················· ····················
X 1820 hCoV-19/Denmark/DCGC-14690/2020|EPI_ISL_711260|202 ···················· ···················· ····················· ···················· ···················· ····················
X 1821 hCoV-19/Denmark/DCGC-14697/2020|EPI_ISL_711261|202 ···················· ···················· ····················· ···················· ···················· ····················
X 1822 hCoV-19/Denmark/DCGC-14708/2020|EPI_ISL_711262|202 ···················· ···················· ····················· ···················· ···················· ····················
X 1823 hCoV-19/Denmark/DCGC-14719/2020|EPI_ISL_711263|202 ···················· ···················· ····················· ···················· ···················· ····················
X 1824 hCoV-19/Denmark/DCGC-14829/2020|EPI_ISL_711264|202 ···················· ···················· ····················· ···················· ···················· ····················
X 1825 hCoV-19/Denmark/DCGC-14833/2020|EPI_ISL_711265|202 ···················· ···················· ····················· ···················· ···················· ····················
X 1826 hCoV-19/Denmark/DCGC-14414/2020|EPI_ISL_711266|202 ···················· ·A·················· ····················· ···················· ···················· ···A················
X 1827 hCoV-19/Denmark/DCGC-14510/2020|EPI_ISL_711267|202 ···················· ·A·················· ····················· ···················· ···················· ···A················
X 1828 hCoV-19/Denmark/DCGC-14869/2020|EPI_ISL_711268|202 ···················· ···················· ····················· ···················· ···················· ····················
X 1829 hCoV-19/Denmark/DCGC-14075/2020|EPI_ISL_711269|202 ···················· ···················· ····················· ···················· ···················· ····················
X 1830 hCoV-19/Denmark/DCGC-14206/2020|EPI_ISL_711270|202 ···················· ···················· ····················· ···················· ···················· ····················
X 1831 hCoV-19/Denmark/DCGC-14907/2020|EPI_ISL_711271|202 ···················· ···················· ····················· ···················· ···················· ····················
X 1832 hCoV-19/Denmark/DCGC-14737/2020|EPI_ISL_711272|202 ···················· ···················· ····················· ···················· ···················· ····················
X 1833 hCoV-19/Denmark/DCGC-14023/2020|EPI_ISL_711286|202 ···················· ···················· ····················· ···················· ···················· ····················
X 1834 hCoV-19/Denmark/DCGC-14076/2020|EPI_ISL_711287|202 ···················· ···················· ····················· ···················· ···················· ····················
X 1835 hCoV-19/Denmark/DCGC-14145/2020|EPI_ISL_711288|202 ···················· ···················· ····················· ···················· ···················· ····················
X 1836 hCoV-19/Denmark/DCGC-14162/2020|EPI_ISL_711289|202 ···················· ···················· ····················· ···················· ···················· ····················
X 1837 hCoV-19/Denmark/DCGC-14237/2020|EPI_ISL_711290|202 ···················· ···················· ····················· ···················· ···················· ····················
X 1838 hCoV-19/Denmark/DCGC-14292/2020|EPI_ISL_711291|202 ···················· ···················· ····················· ···················· ···················· ····················
X 1839 hCoV-19/Denmark/DCGC-14379/2020|EPI_ISL_711292|202 ···················· ···················· ····················· ···················· ···················· ····················
X 1840 hCoV-19/Denmark/DCGC-14392/2020|EPI_ISL_711293|202 ···················· ···················· ····················· ···················· ···················· ····················
X 1841 hCoV-19/Denmark/DCGC-14565/2020|EPI_ISL_711294|202 ···················· ···················· ····················· ···················· ···················· ····················
X 1842 hCoV-19/Denmark/DCGC-14578/2020|EPI_ISL_711295|202 ···················· ···················· ····················· ···················· ···················· ····················
X 1843 hCoV-19/Denmark/DCGC-14638/2020|EPI_ISL_711296|202 ···················· ···················· ····················· ···················· ···················· ····················
X 1844 hCoV-19/Denmark/DCGC-14649/2020|EPI_ISL_711297|202 ···················· ···················· ····················· ···················· ···················· ····················
X 1845 hCoV-19/Denmark/DCGC-14713/2020|EPI_ISL_711298|202 ···················· ···················· ····················· ···················· ···················· ····················
X 1846 hCoV-19/Denmark/DCGC-13011/2020|EPI_ISL_712527|202 ···················· ···················· ····················· ···················· ···················· ····················
X 1847 hCoV-19/Denmark/DCGC-13012/2020|EPI_ISL_712528|202 ···················· ···················· ····················· ···················· ···················· ····················

X 1848 hCoV-19/Denmark/DCGC-13014/2020|EPI_ISL_712529|202 ···················· ···················· ····················· ···················· ···················· ····················
X 1849 hCoV-19/Denmark/DCGC-13015/2020|EPI_ISL_712530|202 ···················· ···················· ····················· ···················· ···················· ····················
X 1850 hCoV-19/Denmark/DCGC-13016/2020|EPI_ISL_712531|202 ···················· ···················· ····················· ···················· ···················· ····················
X 1851 hCoV-19/Denmark/DCGC-13018/2020|EPI_ISL_712532|202 ···················· ···················· ····················· ···················· ···················· ····················
X 1852 hCoV-19/Denmark/DCGC-13019/2020|EPI_ISL_712533|202 ···················· ···················· ····················· ···················· ···················· ····················
X 1853 hCoV-19/Denmark/DCGC-13020/2020|EPI_ISL_712534|202 ···················· ···················· ····················· ···················· ···················· ····················
X 1854 hCoV-19/Denmark/DCGC-13023/2020|EPI_ISL_712535|202 ···················· ···················· ····················· ···················· ···················· ····················
X 1855 hCoV-19/Denmark/DCGC-13024/2020|EPI_ISL_712536|202 ···················· ···················· ····················· ···················· ···················· ····················
X 1856 hCoV-19/Denmark/DCGC-13026/2020|EPI_ISL_712537|202 ···················· ···················· ····················· ···················· ···················· ····················
X 1857 hCoV-19/Denmark/DCGC-13027/2020|EPI_ISL_712538|202 ···················· ···················· ····················· ···················· ···················· ····················
X 1858 hCoV-19/Denmark/DCGC-13028/2020|EPI_ISL_712539|202 ···················· ···················· ····················· ···················· ···················· ····················
X 1859 hCoV-19/Denmark/DCGC-13030/2020|EPI_ISL_712540|202 ···················· ···················· ····················· ···················· ···················· ····················
X 1860 hCoV-19/Denmark/DCGC-13031/2020|EPI_ISL_712541|202 ···················· ···················· ····················· ···················· ···················· ····················
X 1861 hCoV-19/Denmark/DCGC-13032/2020|EPI_ISL_712542|202 ···················· ···················· ····················· ···················· ···················· ····················
X 1862 hCoV-19/Denmark/DCGC-13033/2020|EPI_ISL_712543|202 ···················· ···················· ····················· ···················· ···················· ····················
X 1863 hCoV-19/Denmark/DCGC-13034/2020|EPI_ISL_712544|202 ···················· ···················· ····················· ···················· ···················· ····················
X 1864 hCoV-19/Denmark/DCGC-13035/2020|EPI_ISL_712545|202 ···················· ···················· ····················· ···················· ···················· ····················
X 1865 hCoV-19/Denmark/DCGC-13036/2020|EPI_ISL_712546|202 ···················· ···················· ····················· ···················· ···················· ····················
X 1866 hCoV-19/Denmark/DCGC-13037/2020|EPI_ISL_712547|202 ···················· ···················· ····················· ···················· ···················· ····················
X 1867 hCoV-19/Denmark/DCGC-13038/2020|EPI_ISL_712548|202 ···················· ···················· ····················· ···················· ···················· ····················
X 1868 hCoV-19/Denmark/DCGC-13043/2020|EPI_ISL_712550|202 ···················· ·A·················· ····················· ···················· ···················· ··A·················
X 1869 hCoV-19/Denmark/DCGC-13044/2020|EPI_ISL_712551|202 ···················· ···················· ····················· ···················· ···················· ····················
X 1870 hCoV-19/Denmark/DCGC-13047/2020|EPI_ISL_712553|202 ···················· ···················· ····················· ···················· ···················· ····················
X 1871 hCoV-19/Denmark/DCGC-13061/2020|EPI_ISL_712560|202 ···················· ···················· ····················· ···················· ···················· ····················
X 1872 hCoV-19/Denmark/DCGC-13062/2020|EPI_ISL_712561|202 ···················· ···················· ····················· ···················· ···················· ····················
X 1873 hCoV-19/Denmark/DCGC-13064/2020|EPI_ISL_712562|202 ···················· ···················· ····················· ···················· ···················· ····················
X 1874 hCoV-19/Denmark/DCGC-13065/2020|EPI_ISL_712563|202 ···················· ···················· ····················· ···················· ···················· ····················
X 1875 hCoV-19/Denmark/DCGC-13068/2020|EPI_ISL_712564|202 ···················· ···················· ····················· ···················· ···················· ····················
X 1876 hCoV-19/Denmark/DCGC-13069/2020|EPI_ISL_712565|202 ···················· ···················· ····················· ···················· ···················· ····················
X 1877 hCoV-19/Denmark/DCGC-13070/2020|EPI_ISL_712566|202 ···················· ···················· ····················· ···················· ···················· ····················
X 1878 hCoV-19/Denmark/DCGC-13075/2020|EPI_ISL_712567|202 ···················· ···················· ····················· ···················· ···················· ····················
X 1879 hCoV-19/Tunisia/ADAGE-17149/2020|EPI_ISL_712568|20 ···················· ·A·················· ····················· ···················· ···················· ··A·················
X 1880 hCoV-19/Denmark/DCGC-13076/2020|EPI_ISL_712569|202 ···················· ···················· ····················· ···················· ···················· ····················
X 1881 hCoV-19/Denmark/DCGC-13081/2020|EPI_ISL_712570|202 ···················· ···················· ····················· ···················· ···················· ····················
X 1882 hCoV-19/Denmark/DCGC-13082/2020|EPI_ISL_712571|202 ···················· ···················· ····················· ···················· ···················· ····················
X 1883 hCoV-19/Denmark/DCGC-13083/2020|EPI_ISL_712572|202 ···················· ···················· ····················· ···················· ···················· ····················
X 1884 hCoV-19/Denmark/DCGC-13084/2020|EPI_ISL_712573|202 ···················· ···················· ····················· ···················· ···················· ····················
X 1885 hCoV-19/Denmark/DCGC-13086/2020|EPI_ISL_712574|202 ···················· ···················· ····················· ···················· ···················· ····················
X 1886 hCoV-19/Denmark/DCGC-13087/2020|EPI_ISL_712575|202 ···················· ···················· ····················· ···················· ···················· ····················
X 1887 hCoV-19/Denmark/DCGC-13088/2020|EPI_ISL_712576|202 ···················· ···················· ····················· ···················· ···················· ····················
X 1888 hCoV-19/Denmark/DCGC-13091/2020|EPI_ISL_712577|202 ···················· ···················· ····················· ···················· ···················· ····················
X 1889 hCoV-19/Denmark/DCGC-13094/2020|EPI_ISL_712578|202 ···················· ···················· ····················· ···················· ···················· ····················
X 1890 hCoV-19/Denmark/DCGC-13096/2020|EPI_ISL_712579|202 ···················· ···················· ····················· ···················· ···················· ····················
X 1891 hCoV-19/Denmark/DCGC-13099/2020|EPI_ISL_712580|202 ···················· ···················· ····················· ···················· ···················· ····················
X 1892 hCoV-19/Denmark/DCGC-13100/2020|EPI_ISL_712581|202 ···················· ···················· ····················· ···················· ···················· ····················
X 1893 hCoV-19/Denmark/DCGC-13102/2020|EPI_ISL_712582|202 ···················· ···················· ····················· ···················· ···················· ····················
X 1894 hCoV-19/Denmark/DCGC-13103/2020|EPI_ISL_712583|202 ···················· ···················· ····················· ···················· ···················· ····················
X 1895 hCoV-19/Denmark/DCGC-13104/2020|EPI_ISL_712584|202 ···················· ···················· ····················· ···················· ···················· ····················
X 1896 hCoV-19/Denmark/DCGC-13105/2020|EPI_ISL_712585|202 ···················· ···················· ····················· ···················· ···················· ····················
X 1897 hCoV-19/Denmark/DCGC-13108/2020|EPI_ISL_712586|202 ···················· ···················· ····················· ···················· ···················· ····················
X 1898 hCoV-19/Denmark/DCGC-13109/2020|EPI_ISL_712587|202 ···················· ···················· ····················· ···················· ···················· ····················
X 1899 hCoV-19/Denmark/DCGC-13110/2020|EPI_ISL_712588|202 ···················· ···················· ····················· ···················· ···················· ····················
X 1900 hCoV-19/Denmark/DCGC-13111/2020|EPI_ISL_712589|202 ···················· ···················· ····················· ···················· ···················· ····················
X 1901 hCoV-19/Denmark/DCGC-13112/2020|EPI_ISL_712590|202 ···················· ···················· ····················· ···················· ···················· ····················
X 1902 hCoV-19/Denmark/DCGC-13116/2020|EPI_ISL_712591|202 ···················· ···················· ····················· ···················· ···················· ····················
X 1903 hCoV-19/Denmark/DCGC-13117/2020|EPI_ISL_712592|202 ···················· ···················· ····················· ···················· ···················· ····················
X 1904 hCoV-19/Denmark/DCGC-13118/2020|EPI_ISL_712593|202 ···················· ···················· ····················· ···················· ···················· ····················
X 1905 hCoV-19/Denmark/DCGC-13119/2020|EPI_ISL_712594|202 ···················· ···················· ····················· ···················· ···················· ····················
X 1906 hCoV-19/Denmark/DCGC-13120/2020|EPI_ISL_712595|202 ···················· ···················· ····················· ···················· ···················· ····················
X 1907 hCoV-19/Denmark/DCGC-13122/2020|EPI_ISL_712596|202 ···················· ···················· ····················· ···················· ···················· ····················
X 1908 hCoV-19/Denmark/DCGC-13123/2020|EPI_ISL_712597|202 ···················· ···················· ····················· ···················· ···················· ····················
X 1909 hCoV-19/Denmark/DCGC-13124/2020|EPI_ISL_712598|202 ···················· ···················· ····················· ···················· ···················· ····················
X 1910 hCoV-19/Denmark/DCGC-13126/2020|EPI_ISL_712599|202 ···················· ···················· ····················· ···················· ···················· ····················
X 1911 hCoV-19/Denmark/DCGC-15508/2020|EPI_ISL_713858|202 ···················· ···················· ····················· ···················· ···················· ····················
X 1912 hCoV-19/Denmark/DCGC-15509/2020|EPI_ISL_713859|202 ···················· ···················· ····················· ···················· ···················· ····················
X 1913 hCoV-19/Denmark/DCGC-15511/2020|EPI_ISL_713860|202 ···················· ···················· ····················· ···················· ···················· ····················
X 1914 hCoV-19/Denmark/DCGC-15512/2020|EPI_ISL_713861|202 ···················· ···················· ····················· ···················· ···················· ····················
X 1915 hCoV-19/Denmark/DCGC-15513/2020|EPI_ISL_713862|202 ···················· ···················· ····················· ···················· ···················· ····················
X 1916 hCoV-19/Denmark/DCGC-15516/2020|EPI_ISL_713863|202 ···················· ···················· ····················· ···················· ···················· ····················
X 1917 hCoV-19/Denmark/DCGC-15517/2020|EPI_ISL_713864|202 ···················· ···················· ····················· ···················· ···················· ····················
X 1918 hCoV-19/Denmark/DCGC-15521/2020|EPI_ISL_713865|202 ···················· ···················· ····················· ···················· ···················· ····················
X 1919 hCoV-19/Denmark/DCGC-15522/2020|EPI_ISL_713866|202 ···················· ···················· ····················· ···················· ···················· ····················
X 1920 hCoV-19/Denmark/DCGC-15523/2020|EPI_ISL_713867|202 ···················· ···················· ····················· ···················· ···················· ····················
X 1921 hCoV-19/Denmark/DCGC-15524/2020|EPI_ISL_713868|202 ···················· ···················· ····················· ···················· ···················· ····················
X 1922 hCoV-19/Denmark/DCGC-15525/2020|EPI_ISL_713869|202 ···················· ···················· ····················· ···················· ···················· ····················
X 1923 hCoV-19/Denmark/DCGC-15526/2020|EPI_ISL_713870|202 ···················· ···················· ····················· ···················· ···················· ····················
X 1924 hCoV-19/Denmark/DCGC-15528/2020|EPI_ISL_713871|202 ···················· ···················· ····················· ···················· ···················· ····················
X 1925 hCoV-19/Denmark/DCGC-15529/2020|EPI_ISL_713872|202 ···················· ···················· ····················· ···················· ···················· ····················
X 1926 hCoV-19/Denmark/DCGC-15532/2020|EPI_ISL_713873|202 ···················· ···················· ····················· ···················· ···················· ····················
X 1927 hCoV-19/Denmark/DCGC-15533/2020|EPI_ISL_713874|202 ···················· ···················· ····················· ···················· ···················· ····················

| | | | | | | | |
|---|---|---|---|---|---|---|---|
| X | 1928 hCoV-19/Denmark/DCGC-15534/2020\|EPI_ISL_713875\|202 | ···················· | ···················· | ···················· | ···················· | ···················· | ···················· |
| X | 1929 hCoV-19/Denmark/DCGC-15535/2020\|EPI_ISL_713876\|202 | ···················· | ···················· | ···················· | ···················· | ···················· | ···················· |
| X | 1930 hCoV-19/Denmark/DCGC-15536/2020\|EPI_ISL_713877\|202 | ···················· | ···················· | ···················· | ···················· | ···················· | ···················· |
| X | 1931 hCoV-19/Denmark/DCGC-15537/2020\|EPI_ISL_713878\|202 | ···················· | ···················· | ···················· | ···················· | ···················· | ···················· |
| X | 1932 hCoV-19/Denmark/DCGC-15538/2020\|EPI_ISL_713879\|202 | ···················· | ···················· | ···················· | ···················· | ···················· | ···················· |
| X | 1933 hCoV-19/Denmark/DCGC-15544/2020\|EPI_ISL_713880\|202 | ···················· | ·A·················· | ···················· | ···················· | ···················· | ···A················ |
| X | 1934 hCoV-19/Denmark/DCGC-15545/2020\|EPI_ISL_713881\|202 | ···················· | ···················· | ···················· | ···················· | ···················· | ···················· |
| X | 1935 hCoV-19/Denmark/DCGC-15546/2020\|EPI_ISL_713882\|202 | ···················· | ···················· | ···················· | ···················· | ···················· | ···················· |
| X | 1936 hCoV-19/Denmark/DCGC-15557/2020\|EPI_ISL_713890\|202 | ···················· | ···················· | ···················· | ···················· | ···················· | ···················· |
| X | 1937 hCoV-19/Denmark/DCGC-15558/2020\|EPI_ISL_713891\|202 | ···················· | ···················· | ···················· | ···················· | ···················· | ···················· |
| X | 1938 hCoV-19/Denmark/DCGC-15559/2020\|EPI_ISL_713892\|202 | ···················· | ···················· | ···················· | ···················· | ···················· | ···················· |
| X | 1939 hCoV-19/Denmark/DCGC-15560/2020\|EPI_ISL_713893\|202 | ···················· | ···················· | ···················· | ···················· | ···················· | ···················· |
| X | 1940 hCoV-19/Denmark/DCGC-15561/2020\|EPI_ISL_713894\|202 | ···················· | ···················· | ···················· | ···················· | ···················· | ···················· |
| X | 1941 hCoV-19/Denmark/DCGC-15562/2020\|EPI_ISL_713895\|202 | ···················· | ···················· | ···················· | ···················· | ···················· | ···················· |
| X | 1942 hCoV-19/Denmark/DCGC-15563/2020\|EPI_ISL_713896\|202 | ···················· | ···················· | ···················· | ···················· | ···················· | ···················· |
| X | 1943 hCoV-19/Denmark/DCGC-15565/2020\|EPI_ISL_713897\|202 | ···················· | ···················· | ···················· | ···················· | ···················· | ···················· |
| X | 1944 hCoV-19/Denmark/DCGC-15566/2020\|EPI_ISL_713898\|202 | ···················· | ···················· | ···················· | ···················· | ···················· | ···················· |
| X | 1945 hCoV-19/Denmark/DCGC-15568/2020\|EPI_ISL_713899\|202 | ···················· | ···················· | ···················· | ···················· | ···················· | ···················· |
| X | 1946 hCoV-19/England/ALDP-C48176/2020\|EPI_ISL_718700\|20 | ···················· | ···················· | ···················· | ···················· | ···················· | ···················· |
| X | 1947 hCoV-19/England/CAMC-C47256/2020\|EPI_ISL_718702\|20 | ···················· | ···················· | ···················· | ···················· | ···················· | ···················· |
| X | 1948 hCoV-19/England/CAMC-C478BE/2020\|EPI_ISL_718703\|20 | ···················· | ···················· | ···················· | ···················· | ···················· | ···················· |
| X | 1949 hCoV-19/England/CAMC-C46DCF/2020\|EPI_ISL_718706\|20 | ···················· | ···················· | ···················· | ···················· | ······T············· | ···················· |
| X | 1950 hCoV-19/England/ALDP-C49852/2020\|EPI_ISL_718707\|20 | ···················· | ···················· | ···················· | ···················· | ···················· | ···················· |
| X | 1951 hCoV-19/England/ALDP-C47FD7/2020\|EPI_ISL_718735\|20 | ···················· | ···················· | ···················· | ···················· | ···················· | ···················· |
| X | 1952 hCoV-19/England/ALDP-C47D19/2020\|EPI_ISL_718738\|20 | ···················· | ···················· | ···················· | ···················· | ···················· | ···················· |
| X | 1953 hCoV-19/England/ALDP-C47D28/2020\|EPI_ISL_718739\|20 | ···················· | ···················· | ···················· | ···················· | ···················· | ···················· |
| X | 1954 hCoV-19/England/ALDP-C47D91/2020\|EPI_ISL_718740\|20 | ···················· | ···················· | ···················· | ···················· | ···················· | ···················· |
| X | 1955 hCoV-19/England/ALDP-C47ECB/2020\|EPI_ISL_718744\|20 | ···················· | ···················· | ···················· | ···················· | ···················· | ···················· |
| X | 1956 hCoV-19/England/ALDP-C49C0B/2020\|EPI_ISL_718745\|20 | ···················· | ···················· | ···················· | ···················· | ···················· | ···················· |
| X | 1957 hCoV-19/Netherlands/NH-RIVM-20009/2020\|EPI_ISL_723 | ···················· | ···················· | ···················· | ···················· | ···················· | ···················· |
| X | 1958 hCoV-19/Netherlands/NH-RIVM-20010/2020\|EPI_ISL_723 | ···················· | ···················· | ···················· | ···················· | ···················· | ···················· |
| X | 1959 hCoV-19/Netherlands/NH-RIVM-20011/2020\|EPI_ISL_723 | ···················· | ···················· | ···················· | ···················· | ···················· | ···················· |
| X | 1960 hCoV-19/Netherlands/UT-RIVM-20012/2020\|EPI_ISL_723 | ···················· | ···················· | ···················· | ···················· | ···················· | ···················· |
| X | 1961 hCoV-19/Netherlands/ZE-RIVM-20013/2020\|EPI_ISL_723 | ···················· | ···················· | ···················· | ···················· | ···················· | ···················· |
| X | 1962 hCoV-19/Netherlands/NH-RIVM-20014/2020\|EPI_ISL_723 | ···················· | ···················· | ···················· | ···················· | ···················· | ···················· |
| X | 1963 hCoV-19/Netherlands/NH-RIVM-20015/2020\|EPI_ISL_723 | ···················· | ···················· | ···················· | ···················· | ···················· | ···················· |
| X | 1964 hCoV-19/Netherlands/NH-RIVM-20016/2020\|EPI_ISL_723 | ···················· | ···················· | ···················· | ···················· | ···················· | ···················· |
| X | 1965 hCoV-19/Netherlands/NH-RIVM-20017/2020\|EPI_ISL_723 | ···················· | ···················· | ···················· | ···················· | ···················· | ···················· |
| X | 1966 hCoV-19/Netherlands/NH-RIVM-20018/2020\|EPI_ISL_723 | ···················· | ···················· | ···················· | ···················· | ···················· | ···················· |
| X | 1967 hCoV-19/Netherlands/ZH-RIVM-20019/2020\|EPI_ISL_723 | ···················· | ···················· | ···················· | ···················· | ······T············· | ···················· |
| X | 1968 hCoV-19/Netherlands/NH-RIVM-20020/2020\|EPI_ISL_723 | ···················· | ···················· | ···················· | ···················· | ···················· | ···················· |
| X | 1969 hCoV-19/Netherlands/NH-RIVM-20021/2020\|EPI_ISL_723 | ···················· | ···················· | ···················· | ···················· | ···················· | ···················· |
| X | 1970 hCoV-19/Netherlands/ZH-RIVM-20022/2020\|EPI_ISL_723 | ···················· | ···················· | ···················· | ···················· | ······T············· | ···················· |
| X | 1971 hCoV-19/Netherlands/LI-RIVM-20023/2020\|EPI_ISL_723 | ···················· | ···················· | ···················· | ···················· | ···················· | ···················· |
| X | 1972 hCoV-19/Netherlands/GR-RIVM-20024/2020\|EPI_ISL_723 | ···················· | ···················· | ···················· | ···················· | ···················· | ···················· |
| X | 1973 hCoV-19/Netherlands/NB-RIVM-20025/2020\|EPI_ISL_723 | ···················· | ···················· | ···················· | ···················· | ···················· | ···················· |
| X | 1974 hCoV-19/Netherlands/NB-RIVM-20026/2020\|EPI_ISL_723 | ···················· | ···················· | ···················· | ···················· | ···················· | ···················· |
| X | 1975 hCoV-19/Netherlands/NH-RIVM-20029/2020\|EPI_ISL_723 | ···················· | ···················· | ···················· | ···················· | ···················· | ···················· |
| X | 1976 hCoV-19/Netherlands/NH-RIVM-20030/2020\|EPI_ISL_723 | ···················· | ···················· | ···················· | ···················· | ···················· | ···················· |
| X | 1977 hCoV-19/Netherlands/LI-RIVM-20031/2020\|EPI_ISL_723 | ···················· | ···················· | ···················· | ···················· | ···················· | ···················· |
| X | 1978 hCoV-19/Netherlands/UT-RIVM-20035/2020\|EPI_ISL_723 | ···················· | ···················· | ···················· | ···················· | ···················· | ···················· |
| X | 1979 hCoV-19/Netherlands/UT-RIVM-20036/2020\|EPI_ISL_723 | ···················· | ···················· | ···················· | ···················· | ···················· | ···················· |
| X | 1980 hCoV-19/Netherlands/NH-RIVM-20038/2020\|EPI_ISL_723 | ···················· | ···················· | ···················· | ···················· | ···················· | ···················· |
| X | 1981 hCoV-19/Netherlands/GE-RIVM-20045/2020\|EPI_ISL_723 | ···················· | ···················· | ···················· | ···················· | ···················· | ···················· |
| X | 1982 hCoV-19/Netherlands/NH-RIVM-20048/2020\|EPI_ISL_723 | ···················· | ···················· | ···················· | ···················· | ···················· | ···················· |
| X | 1983 hCoV-19/Netherlands/NH-RIVM-20049/2020\|EPI_ISL_723 | ···················· | ···················· | ···················· | ···················· | ···················· | ···················· |
| X | 1984 hCoV-19/Netherlands/NH-RIVM-20050/2020\|EPI_ISL_723 | ···················· | ···················· | ···················· | ···················· | ···················· | ···················· |
| X | 1985 hCoV-19/Netherlands/NH-RIVM-20052/2020\|EPI_ISL_723 | ···················· | ···················· | ···················· | ···················· | ···················· | ···················· |
| X | 1986 hCoV-19/Netherlands/NH-RIVM-20053/2020\|EPI_ISL_723 | ···················· | ···················· | ···················· | ···················· | ···················· | ···················· |
| X | 1987 hCoV-19/Netherlands/NH-RIVM-20054/2020\|EPI_ISL_723 | ···················· | ···················· | ···················· | ···················· | ···················· | ···················· |
| X | 1988 hCoV-19/Netherlands/UT-RIVM-20055/2020\|EPI_ISL_723 | ···················· | ···················· | ···················· | ···················· | ···················· | ···················· |
| X | 1989 hCoV-19/Netherlands/UT-RIVM-20056/2020\|EPI_ISL_723 | ···················· | ···················· | ···················· | ···················· | ···················· | ···················· |
| X | 1990 hCoV-19/Netherlands/NH-RIVM-20057/2020\|EPI_ISL_723 | ···················· | ···················· | ···················· | ···················· | ···················· | ···················· |
| X | 1991 hCoV-19/Netherlands/NH-RIVM-20058/2020\|EPI_ISL_723 | ···················· | ···················· | ···················· | ···················· | ···················· | ···················· |
| X | 1992 hCoV-19/Netherlands/NH-RIVM-20059/2020\|EPI_ISL_723 | ···················· | ···················· | ···················· | ···················· | ···················· | ···················· |
| X | 1993 hCoV-19/Netherlands/NH-RIVM-20060/2020\|EPI_ISL_723 | ···················· | ···················· | ···················· | ···················· | ···················· | ···················· |
| X | 1994 hCoV-19/Netherlands/NH-RIVM-20061/2020\|EPI_ISL_723 | ···················· | ···················· | ···················· | ···················· | ···················· | ···················· |
| X | 1995 hCoV-19/Netherlands/NH-RIVM-20062/2020\|EPI_ISL_723 | ···················· | ···················· | ···················· | ···················· | ···················· | ···················· |
| X | 1996 hCoV-19/Netherlands/NH-RIVM-20063/2020\|EPI_ISL_723 | ···················· | ···················· | ···················· | ···················· | ···················· | ···················· |
| X | 1997 hCoV-19/Netherlands/NH-RIVM-20064/2020\|EPI_ISL_723 | ···················· | ···················· | ···················· | ···················· | ···················· | ···················· |
| X | 1998 hCoV-19/Netherlands/UT-RIVM-20066/2020\|EPI_ISL_723 | ···················· | ···················· | ···················· | ···················· | ···················· | ···················· |
| X | 1999 hCoV-19/Netherlands/UT-RIVM-20067/2020\|EPI_ISL_723 | ···················· | ···················· | ···················· | ···················· | ···················· | ···················· |
| X | 2000 hCoV-19/Netherlands/GE-RIVM-20069/2020\|EPI_ISL_723 | ···················· | ···················· | ···················· | ···················· | ···················· | ···················· |
| X | 2001 hCoV-19/Netherlands/FR-RIVM-20070/2020\|EPI_ISL_723 | ···················· | ···················· | ···················· | ···················· | ···················· | ···················· |
| X | 2002 hCoV-19/Netherlands/FL-RIVM-20074/2020\|EPI_ISL_723 | ···················· | ···················· | ···················· | ···················· | ···················· | ···················· |
| X | 2003 hCoV-19/Netherlands/NH-RIVM-20075/2020\|EPI_ISL_723 | ···················· | ···················· | ···················· | ···················· | ······T············· | ···················· |
| X | 2004 hCoV-19/Netherlands/NH-RIVM-20076/2020\|EPI_ISL_723 | ···················· | ···················· | ···················· | ···················· | ···················· | ···················· |
| X | 2005 hCoV-19/Netherlands/NH-RIVM-20077/2020\|EPI_ISL_723 | ···················· | ···················· | ···················· | ···················· | ···················· | ···················· |
| X | 2006 hCoV-19/Netherlands/NH-RIVM-20078/2020\|EPI_ISL_723 | ···················· | ···················· | ···················· | ···················· | ···················· | ···················· |
| X | 2007 hCoV-19/Netherlands/NH-RIVM-20079/2020\|EPI_ISL_723 | ···················· | ···················· | ···················· | ···················· | ···················· | ···················· |

```
X 2008 hCoV-19/Netherlands/NH-RIVM-20080/2020|EPI_ISL_723 ···················· ···················· ····················· ···················· ···················· ····················
X 2009 hCoV-19/Netherlands/NH-RIVM-20081/2020|EPI_ISL_723 ···················· ···················· ····················· ···················· ···················· ····················
X 2010 hCoV-19/Netherlands/NH-RIVM-20082/2020|EPI_ISL_723 ···················· ···················· ····················· ···················· ···················· ····················
X 2011 hCoV-19/Netherlands/NH-RIVM-20084/2020|EPI_ISL_723 ···················· ···················· ····················· ···················· ···················· ····················
X 2012 hCoV-19/Netherlands/NH-RIVM-20085/2020|EPI_ISL_723 ···················· ···················· ····················· ···················· ···················· ····················
X 2013 hCoV-19/Netherlands/NH-RIVM-20086/2020|EPI_ISL_723 ···················· ···················· ····················· ···················· ···················· ····················
X 2014 hCoV-19/Netherlands/NH-RIVM-20087/2020|EPI_ISL_723 ···················· ···················· ····················· ···················· ···················· ····················
X 2015 hCoV-19/Netherlands/NH-RIVM-20088/2020|EPI_ISL_723 ···················· ···················· ····················· ···················· ···················· ····················
X 2016 hCoV-19/Netherlands/NH-RIVM-20089/2020|EPI_ISL_723 ···················· ···················· ····················· ···················· ···················· ····················
X 2017 hCoV-19/Netherlands/NH-RIVM-20090/2020|EPI_ISL_723 ···················· ···················· ····················· ···················· ···················· ····················
X 2018 hCoV-19/Netherlands/NH-RIVM-20091/2020|EPI_ISL_723 ···················· ···················· ····················· ···················· ······T············· ····················
X 2019 hCoV-19/Netherlands/UT-RIVM-20092/2020|EPI_ISL_723 ···················· ···················· ····················· ···················· ···················· ····················
X 2020 hCoV-19/Netherlands/OV-RIVM-20093/2020|EPI_ISL_723 ···················· ···················· ····················· ···················· ···················· ····················
X 2021 hCoV-19/Netherlands/OV-RIVM-20094/2020|EPI_ISL_723 ···················· ···················· ····················· ···················· ···················· ····················
X 2022 hCoV-19/Netherlands/GE-RIVM-20095/2020|EPI_ISL_723 ···················· ···················· ····················· ···················· ···················· ····················
X 2023 hCoV-19/Netherlands/OV-RIVM-20096/2020|EPI_ISL_723 ···················· ···················· ····················· ···················· ···················· ····················
X 2024 hCoV-19/Netherlands/GE-RIVM-20097/2020|EPI_ISL_723 ···················· ···················· ····················· ···················· ···················· ····················
X 2025 hCoV-19/Netherlands/OV-RIVM-20098/2020|EPI_ISL_723 ···················· ···················· ····················· ···················· ···················· ····················
X 2026 hCoV-19/Netherlands/FR-RIVM-20100/2020|EPI_ISL_723 ···················· ···················· ····················· ···················· ···················· ····················
X 2027 hCoV-19/Netherlands/FR-RIVM-20101/2020|EPI_ISL_723 ···················· ···················· ····················· ···················· ···················· ····················
X 2028 hCoV-19/Netherlands/FR-RIVM-20102/2020|EPI_ISL_723 ···················· ···················· ····················· ···················· ···················· ····················
X 2029 hCoV-19/Netherlands/NH-RIVM-20103/2020|EPI_ISL_723 ···················· ···················· ····················· ···················· ······T············· ····················
X 2030 hCoV-19/Netherlands/NH-RIVM-20104/2020|EPI_ISL_723 ···················· ···················· ····················· ···················· ···················· ····················
X 2031 hCoV-19/Netherlands/NH-RIVM-20105/2020|EPI_ISL_723 ···················· ···················· ····················· ···················· ···················· ····················
X 2032 hCoV-19/Netherlands/NH-RIVM-20106/2020|EPI_ISL_723 ···················· ···················· ····················· ···················· ···················· ····················
X 2033 hCoV-19/Netherlands/NH-RIVM-20107/2020|EPI_ISL_723 ···················· ···················· ····················· ···················· ···················· ····················
X 2034 hCoV-19/Netherlands/NH-RIVM-20108/2020|EPI_ISL_723 ···················· ···················· ····················· ···················· ···················· ····················
X 2035 hCoV-19/Netherlands/NH-RIVM-20109/2020|EPI_ISL_723 ···················· ···················· ····················· ···················· ···················· ····················
X 2036 hCoV-19/Netherlands/NH-RIVM-20110/2020|EPI_ISL_723 ···················· ···················· ····················· ···················· ···················· ····················
X 2037 hCoV-19/Netherlands/NH-RIVM-20111/2020|EPI_ISL_723 ···················· ···················· ····················· ···················· ···················· ····················
X 2038 hCoV-19/Netherlands/NH-RIVM-20113/2020|EPI_ISL_723 ···················· ···················· ····················· ···················· ···················· ····················
X 2039 hCoV-19/Netherlands/UT-RIVM-20114/2020|EPI_ISL_723 ···················· ···················· ····················· ···················· ···················· ····················
X 2040 hCoV-19/Netherlands/UT-RIVM-20116/2020|EPI_ISL_723 ···················· ···················· ····················· ···················· ···················· ····················
X 2041 hCoV-19/Netherlands/OV-RIVM-20117/2020|EPI_ISL_723 ···················· ···················· ····················· ···················· ···················· ····················
X 2042 hCoV-19/Netherlands/OV-RIVM-20118/2020|EPI_ISL_723 ···················· ···················· ····················· ···················· ···················· ····················
X 2043 hCoV-19/Netherlands/OV-RIVM-20119/2020|EPI_ISL_723 ···················· ···················· ····················· ···················· ···················· ····················
X 2044 hCoV-19/Netherlands/GE-RIVM-20120/2020|EPI_ISL_723 ···················· ······A············· ····················· ···················· ···················· ····················
X 2045 hCoV-19/Netherlands/OV-RIVM-20121/2020|EPI_ISL_723 ···················· ···················· ····················· ···················· ···················· ····················
X 2046 hCoV-19/Netherlands/GE-RIVM-20122/2020|EPI_ISL_723 ···················· ···················· ····················· ···················· ···················· ····················
X 2047 hCoV-19/Netherlands/OV-RIVM-20123/2020|EPI_ISL_723 ···················· ···················· ····················· ···················· ···················· ····················
X 2048 hCoV-19/Netherlands/OV-RIVM-20124/2020|EPI_ISL_723 ···················· ···················· ····················· ···················· ···················· ····················
X 2049 hCoV-19/Scotland/QEUH-BCCD1D/2020|EPI_ISL_710800|2 ···················· ···················· ····················· ···················· ···················· ····················
X 2050 hCoV-19/Scotland/QEUH-BCCC10/2020|EPI_ISL_710802|2 ···················· ···················· ····················· ···················· ···················· ····················
X 2051 hCoV-19/Scotland/QEUH-BCCC6B/2020|EPI_ISL_710803|2 ···················· ···················· ····················· ···················· ···················· ····················
X 2052 hCoV-19/Scotland/QEUH-BCCD0E/2020|EPI_ISL_710804|2 ···················· ···················· ····················· ···················· ···················· ····················
X 2053 hCoV-19/Scotland/QEUH-BCCBE6/2020|EPI_ISL_710805|2 ···················· ···················· ····················· ···················· ···················· ····················
X 2054 hCoV-19/Scotland/QEUH-BCCC01/2020|EPI_ISL_710807|2 ···················· ···················· ····················· ···················· ···················· ····················
X 2055 hCoV-19/Scotland/QEUH-BCCC89/2020|EPI_ISL_710808|2 ···················· ···················· ····················· ···················· ···················· ····················
X 2056 hCoV-19/Scotland/QEUH-BCCBF5/2020|EPI_ISL_710809|2 ···················· ···················· ····················· ···················· ···················· ····················
X 2057 hCoV-19/Scotland/QEUH-BCCBD7/2020|EPI_ISL_710810|2 ···················· ···················· ····················· ···················· ···················· ····················
X 2058 hCoV-19/Scotland/QEUH-BCCD4A/2020|EPI_ISL_710811|2 ···················· ···················· ····················· ···················· ···················· ····················
X 2059 hCoV-19/Scotland/QEUH-BCCD2C/2020|EPI_ISL_710812|2 ···················· ···················· ····················· ···················· ···················· ····················
X 2060 hCoV-19/Scotland/QEUH-BCCD68/2020|EPI_ISL_710813|2 ···················· ···················· ····················· ···················· ···················· ····················
X 2061 hCoV-19/Scotland/QEUH-BCCBC8/2020|EPI_ISL_710814|2 ···················· ···················· ····················· ···················· ···················· ····················
X 2062 hCoV-19/Scotland/QEUH-BCCC98/2020|EPI_ISL_710815|2 ···················· ···················· ····················· ···················· ···················· ····················
X 2063 hCoV-19/Scotland/QEUH-BCCC7A/2020|EPI_ISL_710818|2 ···················· ···················· ····················· ···················· ···················· ····················
X 2064 hCoV-19/Scotland/QEUH-BCD65D/2020|EPI_ISL_710819|2 ···················· ···················· ····················· ···················· ···················· ····················
X 2065 hCoV-19/Scotland/QEUH-BCCBAA/2020|EPI_ISL_710820|2 ···················· ···················· ····················· ···················· ···················· ····················
X 2066 hCoV-19/Wales/CAMC-BC210C/2020|EPI_ISL_710821|2020 ···················· ···················· ····················· ···················· ···················· ····················
X 2067 hCoV-19/Wales/CAMC-BC1C0C/2020|EPI_ISL_710823|2020 ···················· ···················· ····················· ···················· ···················· ····················
X 2068 hCoV-19/Wales/CAMC-BC00C5/2020|EPI_ISL_710824|2020 ···················· ···················· ····················· ···················· ···················· ····················
X 2069 hCoV-19/Wales/CAMC-BC0960/2020|EPI_ISL_710825|2020 ···················· ···················· ····················· ···················· ···················· ····················
X 2070 hCoV-19/Wales/CAMC-BC0A03/2020|EPI_ISL_710826|2020 ···················· ···················· ····················· ···················· ···················· ····················
X 2071 hCoV-19/Wales/CAMC-BC09E8/2020|EPI_ISL_710827|2020 ···················· ···················· ····················· ···················· ···················· ····················
X 2072 hCoV-19/Wales/CAMC-BC0951/2020|EPI_ISL_710828|2020 ···················· ···················· ····················· ···················· ···················· ····················
X 2073 hCoV-19/Wales/CAMC-BC0EBC/2020|EPI_ISL_710829|2020 ···················· ···················· ····················· ···················· ···················· ····················
X 2074 hCoV-19/Scotland/QEUH-BA2EED/2020|EPI_ISL_710831|2 ···················· ···················· ····················· ···················· ······T············· ····················
X 2075 hCoV-19/Scotland/QEUH-BA308C/2020|EPI_ISL_710832|2 ···················· ···················· ····················· ···················· ···················· ····················
X 2076 hCoV-19/Scotland/QEUH-BA2F08/2020|EPI_ISL_710834|2 ···················· ···················· ····················· ···················· ······T············· ····················
X 2077 hCoV-19/Scotland/CAMC-B99B88/2020|EPI_ISL_710835|2 ···················· ···················· ····················· ···················· ···················· ····················
X 2078 hCoV-19/Wales/CAMC-B08F3D/2020|EPI_ISL_710836|2020 ···················· ···················· ····················· ···················· ···················· ····················
X 2079 hCoV-19/Wales/ALDP-B8542A/2020|EPI_ISL_710837|2020 ···················· ···················· ····················· ···················· ···················· ····················
X 2080 hCoV-19/Scotland/QEUH-B7EAE6/2020|EPI_ISL_710838|2 ···················· ···················· ····················· ···················· ···················· ····················
X 2081 hCoV-19/Scotland/QEUH-B80F00/2020|EPI_ISL_710839|2 ···················· ···················· ····················· ···················· ···················· ····················
X 2082 hCoV-19/Scotland/QEUH-B7F344/2020|EPI_ISL_710840|2 ···················· ···················· ····················· ···················· ···················· ····················
X 2083 hCoV-19/Scotland/QEUH-B7F380/2020|EPI_ISL_710841|2 ···················· ···················· ····················· ···················· ···················· ····················
X 2084 hCoV-19/Wales/QEUH-B7C2C2/2020|EPI_ISL_710843|2020 ···················· ···················· ····················· ···················· ···················· ····················
X 2085 hCoV-19/Scotland/QEUH-B7BF9F/2020|EPI_ISL_710844|2 ···················· ···················· ····················· ···················· ···················· ····················
X 2086 hCoV-19/Scotland/QEUH-B7C0F5/2020|EPI_ISL_710845|2 ···················· ···················· ····················· ···················· ···················· ····················
X 2087 hCoV-19/Scotland/QEUH-B7F308/2020|EPI_ISL_710846|2 ···················· ···················· ····················· ···················· ···················· ····················
```

X 2088 hCoV-19/Wales/QEUH-B7C17A/2020|EPI_ISL_710847|2020 ···················· ···················· ····················· ···················· ···················· ····················
X 2089 hCoV-19/Scotland/QEUH-B7F362/2020|EPI_ISL_710848|2 ···················· ···················· ····················· ···················· ···················· ····················
X 2090 hCoV-19/Scotland/QEUH-B7BFDB/2020|EPI_ISL_710849|2 ···················· ···················· ····················· ···················· ···················· ····················
X 2091 hCoV-19/Scotland/QEUH-B7FD28/2020|EPI_ISL_710850|2 ···················· ···················· ····················· ···················· ···················· ····················
X 2092 hCoV-19/Scotland/QEUH-B7F2DE/2020|EPI_ISL_710851|2 ···················· ···················· ····················· ···················· ···················· ····················
X 2093 hCoV-19/Scotland/QEUH-B7C101/2020|EPI_ISL_710852|2 ···················· ···················· ····················· ···················· ···················· ····················
X 2094 hCoV-19/Scotland/QEUH-B7F2ED/2020|EPI_ISL_710853|2 ···················· ···················· ····················· ···················· ···················· ····················
X 2095 hCoV-19/Scotland/QEUH-B7E433/2020|EPI_ISL_710854|2 ···················· ···················· ····················· ···················· ···················· ····················
X 2096 hCoV-19/Scotland/QEUH-B7F2CF/2020|EPI_ISL_710855|2 ···················· ···················· ····················· ···················· ···················· ····················
X 2097 hCoV-19/Wales/QEUH-B7C3ED/2020|EPI_ISL_710856|2020 ···················· ···················· ····················· ···················· ···················· ····················
X 2098 hCoV-19/Scotland/QEUH-B7F317/2020|EPI_ISL_710857|2 ···················· ···················· ····················· ···················· ···················· ····················
X 2099 hCoV-19/Scotland/QEUH-B7F353/2020|EPI_ISL_710858|2 ···················· ···················· ····················· ···················· ···················· ··A·················
X 2100 hCoV-19/Scotland/QEUH-B7F326/2020|EPI_ISL_710859|2 ···················· ···················· ····················· ···················· ···················· ····················
X 2101 hCoV-19/Scotland/QEUH-B7F335/2020|EPI_ISL_710860|2 ···················· ···················· ····················· ···················· ···················· ····················
X 2102 hCoV-19/Wales/QEUH-B812CA/2020|EPI_ISL_710861|2020 ···················· ···················· ····················· ···················· ···················· ····················
X 2103 hCoV-19/Wales/QEUH-B6C7F3/2020|EPI_ISL_710862|2020 ···················· ···················· ····················· ···················· ···················· ····················
X 2104 hCoV-19/Wales/QEUH-B678B9/2020|EPI_ISL_710863|2020 ···················· ···················· ····················· ···················· ···················· ····················
X 2105 hCoV-19/Wales/QEUH-B6792F/2020|EPI_ISL_710864|2020 ···················· ···················· ····················· ···················· ···················· ····················
X 2106 hCoV-19/Wales/ALDP-B5CF67/2020|EPI_ISL_710865|2020 ···················· ···················· ····················· ···················· ···················· ····················
X 2107 hCoV-19/Wales/ALDP-B5CC33/2020|EPI_ISL_710866|2020 ···················· ···················· ····················· ···················· ···················· ····················
X 2108 hCoV-19/Wales/QEUH-B6C75D/2020|EPI_ISL_710867|2020 ···················· ···················· ····················· ···················· ···················· ····················
X 2109 hCoV-19/Wales/QEUH-B678F5/2020|EPI_ISL_710868|2020 ···················· ···················· ····················· ···················· ···················· ····················
X 2110 hCoV-19/Wales/ALDP-B5CF1C/2020|EPI_ISL_710869|2020 ···················· ···················· ····················· ···················· ···················· ····················
X 2111 hCoV-19/Wales/QEUH-B6C140/2020|EPI_ISL_710872|2020 ···················· ···················· ····················· ···················· ···················· ····················
X 2112 hCoV-19/Wales/ALDP-B6301F/2020|EPI_ISL_710873|2020 ···················· ···················· ····················· ···················· ···················· ····················
X 2113 hCoV-19/Wales/ALDP-B62FF5/2020|EPI_ISL_710874|2020 ···················· ···················· ····················· ···················· ···················· ····················
X 2114 hCoV-19/Wales/ALDP-B630F1/2020|EPI_ISL_710875|2020 ···················· ···················· ····················· ···················· ···················· ····················
X 2115 hCoV-19/Wales/ALDP-B630A6/2020|EPI_ISL_710876|2020 ···················· ···················· ····················· ···················· ···················· ····················
X 2116 hCoV-19/Wales/ALDP-B49264/2020|EPI_ISL_710888|2020 ···················· ···················· ····················· ···················· ···················· ····················
X 2117 hCoV-19/Wales/ALDP-B49228/2020|EPI_ISL_710889|2020 ···················· ···················· ····················· ···················· ···················· ····················
X 2118 hCoV-19/Wales/ALDP-B491C1/2020|EPI_ISL_710890|2020 ···················· ···················· ····················· ···················· ···················· ····················
X 2119 hCoV-19/Wales/ALDP-B47E71/2020|EPI_ISL_710891|2020 ···················· ···················· ····················· ···················· ···················· ····················
X 2120 hCoV-19/Wales/ALDP-B47E80/2020|EPI_ISL_710892|2020 ···················· ···················· ····················· ···················· ···················· ····················
X 2121 hCoV-19/Wales/ALDP-B47D47/2020|EPI_ISL_710893|2020 ···················· ···················· ····················· ···················· ···················· ····················
X 2122 hCoV-19/Wales/ALDP-B47D1A/2020|EPI_ISL_710894|2020 ···················· ···················· ····················· ···················· ···················· ····················
X 2123 hCoV-19/Wales/ALDP-B4870C/2020|EPI_ISL_710895|2020 ···················· ···················· ····················· ···················· ···················· ····················
X 2124 hCoV-19/Wales/ALDP-B492DD/2020|EPI_ISL_710896|2020 ···················· ···················· ····················· ···················· ···················· ····················
X 2125 hCoV-19/Wales/ALDP-B48818/2020|EPI_ISL_710897|2020 ···················· ···················· ····················· ···················· ···················· ····················
X 2126 hCoV-19/Wales/ALDP-B487C0/2020|EPI_ISL_710898|2020 ···················· ···················· ····················· ···················· ···················· ····················
X 2127 hCoV-19/Wales/ALDP-B49291/2020|EPI_ISL_710899|2020 ···················· ···················· ····················· ···················· ···················· ····················
X 2128 hCoV-19/Denmark/DCGC-13851/2020|EPI_ISL_713060|202 ···················· ···················· ····················· ···················· ···················· ····················
X 2129 hCoV-19/Denmark/DCGC-13854/2020|EPI_ISL_713061|202 ···················· ···················· ····················· ···················· ···················· ····················
X 2130 hCoV-19/Denmark/DCGC-13855/2020|EPI_ISL_713062|202 ···················· ···················· ····················· ···················· ···················· ····················
X 2131 hCoV-19/Denmark/DCGC-13856/2020|EPI_ISL_713063|202 ···················· ···················· ····················· ···················· ···················· ····················
X 2132 hCoV-19/Denmark/DCGC-13857/2020|EPI_ISL_713064|202 ···················· ···················· ····················· ···················· ···················· ····················
X 2133 hCoV-19/Denmark/DCGC-13858/2020|EPI_ISL_713065|202 ···················· ···················· ····················· ···················· ···················· ····················
X 2134 hCoV-19/Denmark/DCGC-13859/2020|EPI_ISL_713066|202 ···················· ···················· ····················· ···················· ···················· ····················
X 2135 hCoV-19/Denmark/DCGC-13860/2020|EPI_ISL_713067|202 ···················· ···················· ····················· ···················· ···················· ····················
X 2136 hCoV-19/Denmark/DCGC-13864/2020|EPI_ISL_713068|202 ···················· ···················· ····················· ···················· ···················· ····················
X 2137 hCoV-19/Denmark/DCGC-13866/2020|EPI_ISL_713069|202 ···················· ···················· ····················· ···················· ···················· ····················
X 2138 hCoV-19/Denmark/DCGC-13868/2020|EPI_ISL_713070|202 ···················· ···················· ····················· ···················· ···················· ····················
X 2139 hCoV-19/Denmark/DCGC-13869/2020|EPI_ISL_713071|202 ···················· ···················· ····················· ···················· ···················· ····················
X 2140 hCoV-19/Denmark/DCGC-13870/2020|EPI_ISL_713072|202 ···················· ···················· ····················· ···················· ····T··············· ····················
X 2141 hCoV-19/Denmark/DCGC-13871/2020|EPI_ISL_713073|202 ···················· ·A·················· ····················· ···················· ···················· ····················
X 2142 hCoV-19/Denmark/DCGC-13872/2020|EPI_ISL_713074|202 ···················· ···················· ····················· ···················· ···················· ····················
X 2143 hCoV-19/Denmark/DCGC-13874/2020|EPI_ISL_713075|202 ···················· ···················· ····················· ···················· ···················· ····················
X 2144 hCoV-19/Denmark/DCGC-13875/2020|EPI_ISL_713076|202 ···················· ···················· ····················· ···················· ···················· ····················
X 2145 hCoV-19/Denmark/DCGC-13877/2020|EPI_ISL_713077|202 ···················· ···················· ····················· ···················· ···················· ····················
X 2146 hCoV-19/Denmark/DCGC-13878/2020|EPI_ISL_713078|202 ···················· ···················· ····················· ···················· ···················· ····················
X 2147 hCoV-19/Denmark/DCGC-13879/2020|EPI_ISL_713079|202 ···················· ···················· ····················· ···················· ···················· ····················
X 2148 hCoV-19/Denmark/DCGC-13880/2020|EPI_ISL_713080|202 ···················· ···················· ····················· ···················· ···················· ····················
X 2149 hCoV-19/Denmark/DCGC-13883/2020|EPI_ISL_713081|202 ···················· ···················· ····················· ···················· ···················· ····················
X 2150 hCoV-19/Denmark/DCGC-13884/2020|EPI_ISL_713082|202 ···················· ···················· ····················· ···················· ···················· ····················
X 2151 hCoV-19/Denmark/DCGC-13885/2020|EPI_ISL_713083|202 ···················· ···················· ····················· ···················· ···················· ····················
X 2152 hCoV-19/Denmark/DCGC-13889/2020|EPI_ISL_713084|202 ···················· ···················· ····················· ···················· ···················· ····················
X 2153 hCoV-19/Denmark/DCGC-13892/2020|EPI_ISL_713085|202 ···················· ···················· ····················· ···················· ···················· ····················
X 2154 hCoV-19/Denmark/DCGC-13893/2020|EPI_ISL_713086|202 ···················· ···················· ····················· ···················· ···················· ····················
X 2155 hCoV-19/Denmark/DCGC-13894/2020|EPI_ISL_713087|202 ···················· ···················· ····················· ···················· ···················· ····················
X 2156 hCoV-19/Denmark/DCGC-13895/2020|EPI_ISL_713088|202 ···················· ···················· ····················· ···················· ···················· ····················
X 2157 hCoV-19/Denmark/DCGC-13897/2020|EPI_ISL_713089|202 ···················· ···················· ····················· ···················· ···················· ····················
X 2158 hCoV-19/Denmark/DCGC-13898/2020|EPI_ISL_713090|202 ···················· ···················· ····················· ···················· ···················· ····················
X 2159 hCoV-19/Denmark/DCGC-13899/2020|EPI_ISL_713091|202 ···················· ···················· ··T·················· ···················· ···················· ····················
X 2160 hCoV-19/Denmark/DCGC-13900/2020|EPI_ISL_713092|202 ···················· ···················· ····················· ···················· ···················· ····················
X 2161 hCoV-19/Denmark/DCGC-13902/2020|EPI_ISL_713093|202 ···················· ···················· ····················· ···················· ···················· ····················
X 2162 hCoV-19/Denmark/DCGC-13903/2020|EPI_ISL_713094|202 ···················· ···················· ····················· ···················· ···················· ····················
X 2163 hCoV-19/Denmark/DCGC-13906/2020|EPI_ISL_713095|202 ···················· ···················· ····················· ···················· ···················· ····················
X 2164 hCoV-19/Denmark/DCGC-13907/2020|EPI_ISL_713096|202 ···················· ···················· ····················· ···················· ···················· ····················
X 2165 hCoV-19/Denmark/DCGC-13908/2020|EPI_ISL_713097|202 ···················· ···················· ····················· ···················· ···················· ····················
X 2166 hCoV-19/England/CAMC-C46E35/2020|EPI_ISL_718709|20 ···················· ···················· ····················· ···················· ···················· ····················
X 2167 hCoV-19/England/ALDP-C480D3/2020|EPI_ISL_718711|20 ···················· ···················· ····················· ···················· ···················· ····················

X 2168 hCoV-19/England/ALDP-C49A01/2020|EPI_ISL_718712|20 ···················· ···················· ····················· ···················· ···················· ····················
X 2169 hCoV-19/England/ALDP-C49A2F/2020|EPI_ISL_718713|20 ···················· ···················· ····················· ···················· ···················· ····················
X 2170 hCoV-19/England/ALDP-C48194/2020|EPI_ISL_718714|20 ···················· ···················· ····················· ···················· ···················· ····················
X 2171 hCoV-19/England/ALDP-C49A4D/2020|EPI_ISL_718715|20 ···················· ···················· ····················· ···················· ···················· ····················
X 2172 hCoV-19/England/ALDP-C49A6B/2020|EPI_ISL_718716|20 ···················· ···················· ····················· ···················· ······T············· ····················
X 2173 hCoV-19/England/CAMC-C470F2/2020|EPI_ISL_718717|20 ···················· ···················· ····················· ···················· ···················· ····················
X 2174 hCoV-19/England/ALDP-C48000/2020|EPI_ISL_718718|20 ···················· ···················· ····················· ···················· ···················· ····················
X 2175 hCoV-19/England/ALDP-C47CB2/2020|EPI_ISL_718719|20 ···················· ···················· ····················· ···················· ···················· ····················
X 2176 hCoV-19/England/ALDP-C48097/2020|EPI_ISL_718721|20 ···················· ···················· ····················· ···················· ···················· ····················
X 2177 hCoV-19/England/CAMC-C4739F/2020|EPI_ISL_718722|20 ···················· ···················· ····················· ···················· ···················· ····················
X 2178 hCoV-19/England/ALDP-C47F40/2020|EPI_ISL_718725|20 ···················· ···················· ····················· ···················· ···················· ····················
X 2179 hCoV-19/England/CAMC-C471A4/2020|EPI_ISL_718726|20 ···················· ···················· ····················· ···················· ···················· ····················
X 2180 hCoV-19/England/CAMC-C47177/2020|EPI_ISL_718727|20 ···················· ···················· ····················· ···················· ···················· ····················
X 2181 hCoV-19/England/ALDP-C480A6/2020|EPI_ISL_718728|20 ···················· ···················· ····················· ···················· ···A················ ····················
X 2182 hCoV-19/England/ALDP-C47E52/2020|EPI_ISL_718747|20 ···················· ···················· ····················· ···················· ···················· ····················
X 2183 hCoV-19/England/CAMC-C3E586/2020|EPI_ISL_719205|20 ···················· ···················· ····················· ···················· ·········T·········· ····················
X 2184 hCoV-19/England/CAMC-C43858/2020|EPI_ISL_719221|20 ···················· ···················· ····················· ···················· ···················· ····················
X 2185 hCoV-19/England/CAMC-C3EDF7/2020|EPI_ISL_719226|20 ···················· ···················· ····················· ···················· ···················· ····················
X 2186 hCoV-19/England/CAMC-C43A52/2020|EPI_ISL_719228|20 ···················· ···················· ····················· ···················· ···················· ····················
X 2187 hCoV-19/England/CAMC-C41BAC/2020|EPI_ISL_719230|20 ···················· ···················· ····················· ···················· ···················· ····················
X 2188 hCoV-19/England/CAMC-C43B31/2020|EPI_ISL_719235|20 ···················· ···················· ····················· ···················· ···················· ····················
X 2189 hCoV-19/England/CAMC-C40C31/2020|EPI_ISL_719236|20 ···················· ···················· ····················· ···················· ···················· ····················
X 2190 hCoV-19/England/MILK-BB1C87/2020|EPI_ISL_720205|20 ···················· ···················· ····················· ···················· ···················· ····················
X 2191 hCoV-19/England/MILK-BB1461/2020|EPI_ISL_720206|20 ···················· ···················· ····················· ···················· ······T············· ····················
X 2192 hCoV-19/England/MILK-BB271C/2020|EPI_ISL_720207|20 ···················· ···················· ····················· ···················· ···················· ····················
X 2193 hCoV-19/England/MILK-BB1FBB/2020|EPI_ISL_720208|20 ···················· ···················· ····················· ···················· ···················· ····················
X 2194 hCoV-19/England/MILK-BB1346/2020|EPI_ISL_720210|20 ···················· ···················· ····················· ···················· ···················· ····················
X 2195 hCoV-19/England/MILK-BB01A7/2020|EPI_ISL_720211|20 ···················· ···················· ····················· ···················· ···················· ···A················
X 2196 hCoV-19/England/MILK-BB20D5/2020|EPI_ISL_720214|20 ···················· ···················· ····················· ···················· ···················· ····················
X 2197 hCoV-19/England/MILK-BB0189/2020|EPI_ISL_720215|20 ···················· ···················· ····················· ···················· ···················· ····················
X 2198 hCoV-19/England/MILK-BB264C/2020|EPI_ISL_720217|20 ···················· ···················· ····················· ···················· ······T············· ····················
X 2199 hCoV-19/England/MILK-BB24D9/2020|EPI_ISL_720218|20 ···················· ···················· ····················· ···················· ···················· ····················
X 2200 hCoV-19/England/MILK-BB2503/2020|EPI_ISL_720219|20 ···················· ···················· ····················· ···················· ···················· ····················
X 2201 hCoV-19/England/MILK-BB0A60/2020|EPI_ISL_720221|20 ···················· ···················· ····················· ···················· ···················· ····················
X 2202 hCoV-19/England/MILK-BB08C0/2020|EPI_ISL_720222|20 ···················· ···················· ····················· ···················· ···················· ····················
X 2203 hCoV-19/England/MILK-BB01C5/2020|EPI_ISL_720223|20 ···················· ···················· ····················· ···················· ···················· ···A················
X 2204 hCoV-19/England/ALDP-BC84EF/2020|EPI_ISL_721500|20 ···················· ···················· ····················· ···················· ···················· ····················
X 2205 hCoV-19/England/ALDP-BC750B/2020|EPI_ISL_721501|20 ···················· ···················· ····················· ···················· ···················· ····················
X 2206 hCoV-19/England/ALDP-BC7B63/2020|EPI_ISL_721502|20 ···················· ···················· ····················· ···················· ···················· ····················
X 2207 hCoV-19/England/ALDP-BC7468/2020|EPI_ISL_721503|20 ···················· ···················· ····················· ···················· ···················· ····················
X 2208 hCoV-19/England/ALDP-BC64C3/2020|EPI_ISL_721504|20 ···················· ···················· ····················· ···················· ···················· ····················
X 2209 hCoV-19/England/ALDP-BC86F8/2020|EPI_ISL_721505|20 ···················· ···················· ····················· ···················· ···················· ····················
X 2210 hCoV-19/England/ALDP-BC6830/2020|EPI_ISL_721506|20 ···················· ···················· ····················· ···················· ···················· ····················
X 2211 hCoV-19/England/ALDP-BC779C/2020|EPI_ISL_721507|20 ···················· ···················· ····················· ···················· ···················· ····················
X 2212 hCoV-19/England/ALDP-BC8573/2020|EPI_ISL_721508|20 ···················· ···················· ····················· ···················· ···················· ····················
X 2213 hCoV-19/England/ALDP-BC8670/2020|EPI_ISL_721509|20 ···················· ···················· ····················· ···················· ···················· ····················
X 2214 hCoV-19/England/ALDP-BC876E/2020|EPI_ISL_721510|20 ···················· ···················· ····················· ···················· ···················· ····················
X 2215 hCoV-19/England/ALDP-BC744A/2020|EPI_ISL_721512|20 ···················· ···················· ····················· ···················· ······T············· ····················
X 2216 hCoV-19/England/ALDP-BC784E/2020|EPI_ISL_721513|20 ···················· ···················· ····················· ···················· ···················· ····················
X 2217 hCoV-19/England/ALDP-C06806/2020|EPI_ISL_721518|20 ···················· ···················· ····················· ···················· ···················· ····················
X 2218 hCoV-19/England/ALDP-C06226/2020|EPI_ISL_721520|20 ···················· ···················· ····················· ···················· ···················· ····················
X 2219 hCoV-19/England/QEUH-C17C60/2020|EPI_ISL_721521|20 ···················· ···················· ····················· ···················· ···················· ····················
X 2220 hCoV-19/England/QEUH-A8B7FF/2020|EPI_ISL_721536|20 ···················· ···················· ····················· ···················· ···················· ····················
X 2221 hCoV-19/England/QEUH-A8C2FD/2020|EPI_ISL_721537|20 ···················· ···················· ····················· ···················· ···················· ····················
X 2222 hCoV-19/England/QEUH-A8B80B/2020|EPI_ISL_721538|20 ···················· ···················· ····················· ···················· ···················· ····················
X 2223 hCoV-19/Scotland/QEUH-A8B6A7/2020|EPI_ISL_721540|2 ···················· ···················· ····················· ···················· ···················· ····················
X 2224 hCoV-19/England/QEUH-A8C776/2020|EPI_ISL_721541|20 ···················· ···················· ····················· ···················· ···················· ····················
X 2225 hCoV-19/England/QEUH-AAE204/2020|EPI_ISL_721543|20 ···················· ···················· ····················· ···················· ···················· ····················
X 2226 hCoV-19/England/QEUH-AAE5ED/2020|EPI_ISL_721544|20 ···················· ···················· ····················· ···················· ···················· ····················
X 2227 hCoV-19/England/QEUH-AADD9B/2020|EPI_ISL_721548|20 ···················· ···················· ····················· ···················· ···················· ····················
X 2228 hCoV-19/England/QEUH-AAE1F8/2020|EPI_ISL_721549|20 ···················· ···················· ····················· ···················· ···················· ····················
X 2229 hCoV-19/England/QEUH-AAE213/2020|EPI_ISL_721551|20 ···················· ···················· ····················· ···················· ···················· ····················
X 2230 hCoV-19/England/QEUH-AAF35B/2020|EPI_ISL_721552|20 ···················· ···················· ····················· ···················· ···················· ····················
X 2231 hCoV-19/England/QEUH-AAE143/2020|EPI_ISL_721554|20 ···················· ···················· ····················· ···················· ···················· ····················
X 2232 hCoV-19/Netherlands/ZE-EMC-74/2020|EPI_ISL_722800| ···················· ···················· ····················· ···················· ···················· ····················
X 2233 hCoV-19/Netherlands/NB-EMC-451/2020|EPI_ISL_722801 ···················· ···················· ····················· ···················· ···················· ····················
X 2234 hCoV-19/Netherlands/NB-EMC-452/2020|EPI_ISL_722802 ···················· ···················· ····················· ···················· ···················· ····················
X 2235 hCoV-19/Netherlands/NB-EMC-454/2020|EPI_ISL_722803 ···················· ···················· ····················· ···················· ···················· ····················
X 2236 hCoV-19/Netherlands/NB-EMC-455/2020|EPI_ISL_722804 ···················· ···················· ····················· ···················· ···················· ····················
X 2237 hCoV-19/Netherlands/NB-EMC-456/2020|EPI_ISL_722805 ···················· ···················· ····················· ···················· ···················· ····················
X 2238 hCoV-19/Netherlands/NB-EMC-457/2020|EPI_ISL_722806 ···················· ···················· ····················· ···················· ···················· ····················
X 2239 hCoV-19/Netherlands/NB-EMC-458/2020|EPI_ISL_722807 ···················· ···················· ····················· ···················· ···················· ····················
X 2240 hCoV-19/Netherlands/NB-EMC-459/2020|EPI_ISL_722808 ···················· ···················· ····················· ···················· ···················· ····················
X 2241 hCoV-19/Netherlands/NB-EMC-460/2020|EPI_ISL_722809 ···················· ···················· ····················· ···················· ···················· ····················
X 2242 hCoV-19/Netherlands/NB-EMC-463/2020|EPI_ISL_722810 ···················· ···················· ····················· ···················· ···················· ····················
X 2243 hCoV-19/Netherlands/NB-EMC-464/2020|EPI_ISL_722811 ···················· ···················· ····················· ···················· ···················· ····················
X 2244 hCoV-19/Netherlands/NB-EMC-467/2020|EPI_ISL_722812 ···················· ···················· ····················· ···················· ···················· ····················
X 2245 hCoV-19/Netherlands/NB-EMC-468/2020|EPI_ISL_722813 ···················· ···················· ····················· ···················· ···················· ····················
X 2246 hCoV-19/Netherlands/NB-EMC-469/2020|EPI_ISL_722814 ···················· ···················· ····················· ···················· ···················· ····················
X 2247 hCoV-19/Netherlands/ZH-EMC-1024/2020|EPI_ISL_72281 ···················· ···················· ····················· ···················· ···················· ····················

X 2248 hCoV-19/Netherlands/OV-EMC-91/2020|EPI_ISL_722816| ···················· ···················· ···················· ···················· ···················· ····················
X 2249 hCoV-19/Netherlands/OV-EMC-93/2020|EPI_ISL_722817| ···················· ···················· ···················· ···················· ···················· ····················
X 2250 hCoV-19/Netherlands/GE-EMC-434/2020|EPI_ISL_722818 ···················· ···················· ···················· ···················· ···················· ····················
X 2251 hCoV-19/Netherlands/OV-EMC-95/2020|EPI_ISL_722819| ···················· ···················· ···················· ···················· ···················· ····················
X 2252 hCoV-19/Netherlands/OV-EMC-96/2020|EPI_ISL_722820| ···················· ···················· ···················· ···················· ···················· ····················
X 2253 hCoV-19/Netherlands/OV-EMC-100/2020|EPI_ISL_722821 ···················· ···················· ···················· ···················· ···················· ····················
X 2254 hCoV-19/Netherlands/OV-EMC-102/2020|EPI_ISL_722822 ···················· ···················· ···················· ···················· ···················· ····················
X 2255 hCoV-19/Netherlands/OV-EMC-103/2020|EPI_ISL_722823 ···················· ···················· ···················· ···················· ···················· ····················
X 2256 hCoV-19/Netherlands/OV-EMC-104/2020|EPI_ISL_722824 ···················· ···················· ···················· ···················· ···················· ····················
X 2257 hCoV-19/Netherlands/OV-EMC-105/2020|EPI_ISL_722825 ···················· ···················· ···················· ···················· ···················· ····················
X 2258 hCoV-19/Netherlands/ZH-EMC-991/2020|EPI_ISL_722826 ···················· ···················· ···················· ···················· ···················· ····················
X 2259 hCoV-19/Netherlands/NB-EMC-378/2020|EPI_ISL_722827 ···················· ···················· ···················· ···················· ···················· ····················
X 2260 hCoV-19/Netherlands/ZH-EMC-960/2020|EPI_ISL_722828 ···················· ···················· ···················· ···················· ···················· ····················
X 2261 hCoV-19/Netherlands/NB-EMC-379/2020|EPI_ISL_722829 ···················· ···················· ···················· ···················· ···················· ····················
X 2262 hCoV-19/Netherlands/OV-EMC-106/2020|EPI_ISL_722830 ···················· ···················· ···················· ···················· ···················· ····················
X 2263 hCoV-19/Netherlands/OV-EMC-107/2020|EPI_ISL_722831 ···················· ···················· ···················· ···················· ···················· ····················
X 2264 hCoV-19/Netherlands/NB-EMC-374/2020|EPI_ISL_722832 ···················· ···················· ···················· ···················· ···················· ····················
X 2265 hCoV-19/Netherlands/NB-EMC-375/2020|EPI_ISL_722833 ···················· ···················· ···················· ···················· ···················· ····················
X 2266 hCoV-19/Netherlands/NB-EMC-380/2020|EPI_ISL_722834 ···················· ···················· ···················· ···················· ···················· ····················
X 2267 hCoV-19/Netherlands/NB-EMC-381/2020|EPI_ISL_722835 ···················· ···················· ···················· ···················· ···················· ····················
X 2268 hCoV-19/Netherlands/NB-EMC-382/2020|EPI_ISL_722836 ···················· ···················· ···················· ···················· ···················· ····················
X 2269 hCoV-19/Netherlands/OV-EMC-109/2020|EPI_ISL_722837 ···················· ···················· ···················· ···················· ···················· ····················
X 2270 hCoV-19/Netherlands/GE-EMC-392/2020|EPI_ISL_722838 ···················· ···················· ···················· ···················· ···················· ····················
X 2271 hCoV-19/Netherlands/OV-EMC-108/2020|EPI_ISL_722839 ···················· ···················· ···················· ···················· ···················· ····················
X 2272 hCoV-19/Belgium/un-EMC-6/2020/2020|EPI_ISL_722840| ···················· ···················· ···················· ···················· ···················· ····················
X 2273 hCoV-19/Netherlands/NB-EMC-383/2020|EPI_ISL_722841 ···················· ···················· ···················· ···················· ···················· ····················
X 2274 hCoV-19/Netherlands/OV-EMC-85/2020|EPI_ISL_722842| ···················· ···················· ···················· ···················· ···················· ····················
X 2275 hCoV-19/Netherlands/OV-EMC-84/2020|EPI_ISL_722843| ···················· ···················· ···················· ···················· ···················· ····················
X 2276 hCoV-19/Netherlands/OV-EMC-90/2020|EPI_ISL_722844| ···················· ···················· ···················· ···················· ···················· ····················
X 2277 hCoV-19/Netherlands/ZH-EMC-906/2020|EPI_ISL_722850 ···················· ···················· ···················· ···················· ······T············· ····················
X 2278 hCoV-19/Italy/APU-IZSPB-177APT/2020|EPI_ISL_722851 ···················· ···················· ···················· ···················· ···················· ····················
X 2279 hCoV-19/Italy/APU-IZSPB-179APT/2020|EPI_ISL_722852 ···················· ···················· ···················· ···················· ···················· ····················
X 2280 hCoV-19/Italy/APU-IZSPB-191APT/2020|EPI_ISL_722853 ···················· ···················· ···················· ···················· ···················· ····················
X 2281 hCoV-19/Italy/APU-IZSPB-199APT/2020|EPI_ISL_722854 ···················· ···················· ···················· ···················· ···················· ····················
X 2282 hCoV-19/Italy/APU-IZSPB-147APT/2020|EPI_ISL_722855 ···················· ···················· ···················· ···················· ···················· ····················
X 2283 hCoV-19/Italy/APU-IZSPB-195PT/2020|EPI_ISL_722867| ···················· ···················· ···················· ···················· ···················· ····················
X 2284 hCoV-19/Italy/APU-IZSPB-197PT/2020|EPI_ISL_722868| ···················· ···················· ···················· ···················· ···················· ····················
X 2285 hCoV-19/Italy/APU-IZSPB-201PT/2020|EPI_ISL_722869| ···················· ···················· ···················· ···················· ···················· ····················
X 2286 hCoV-19/Italy/APU-IZSPB-202PT/2020|EPI_ISL_722870| ···················· ···················· ···················· ···················· ···················· ····················
X 2287 hCoV-19/Italy/APU-IZSPB-203PT/2020|EPI_ISL_722871| ···················· ···················· ···················· ···················· ···················· ····················
X 2288 hCoV-19/Italy/APU-IZSPB-158PT/2020|EPI_ISL_722872| ···················· ···················· ···················· ···················· ···················· ····················
X 2289 hCoV-19/Italy/BAS-IZSPB-338PT/2020|EPI_ISL_722873| ···················· ·A·················· ···················· ···················· ···················· ····················
X 2290 hCoV-19/Italy/BAS-IZSPB-339PT/2020|EPI_ISL_722874| ···················· ···················· ···················· ···················· ···················· ····················
X 2291 hCoV-19/Italy/BAS-IZSPB-341PT/2020|EPI_ISL_722875| ···················· ···················· ···················· ···················· ···················· ····················
X 2292 hCoV-19/Italy/BAS-IZSPB-351PT/2020|EPI_ISL_722876| ···················· ···················· ···················· ···················· ···················· ····················
X 2293 hCoV-19/Italy/BAS-IZSPB-359PT/2020|EPI_ISL_722877| ···················· ···················· ···················· ···················· ···················· ····················
X 2294 hCoV-19/Italy/BAS-IZSPB-364PT/2020|EPI_ISL_722878| ···················· ···················· ···················· ···················· ···················· ····················
X 2295 hCoV-19/Italy/BAS-IZSPB-368PT/2020|EPI_ISL_722879| ···················· ···················· ···················· ···················· ···················· ····················
X 2296 hCoV-19/Italy/BAS-IZSPB-373PT/2020|EPI_ISL_722880| ···················· ···················· ···················· ···················· ···················· ····················
X 2297 hCoV-19/Italy/BAS-IZSPB-374PT/2020|EPI_ISL_722881| ···················· ···················· ···················· ···················· ···················· ····················
X 2298 hCoV-19/Italy/BAS-IZSPB-380PT/2020|EPI_ISL_722882| ···················· ···················· ···················· ···················· ···················· ····················
X 2299 hCoV-19/Italy/BAS-IZSPB-381PT/2020|EPI_ISL_722883| ··················T· ···················· ···················· ···················· ······T············· ···················· 1
X 2300 hCoV-19/Italy/BAS-IZSPB-383PT/2020|EPI_ISL_722884| ···················· ···················· ···················· ···················· ···················· ····················
X 2301 hCoV-19/Italy/BAS-IZSPB-384PT/2020|EPI_ISL_722885| ···················· ···················· ···················· ···················· ···················· ····················
X 2302 hCoV-19/Italy/BAS-IZSPB-385PT/2020|EPI_ISL_722886| ···················· ···················· ···················· ···················· ···················· ····················
X 2303 hCoV-19/Italy/BAS-IZSPB-388PT/2020|EPI_ISL_722887| ···················· ···················· ···················· ···················· ···················· ····················
X 2304 hCoV-19/Italy/BAS-IZSPB-389PT/2020|EPI_ISL_722888| ···················· ···················· ···················· ···················· ···················· ····················
X 2305 hCoV-19/Wales/QEUH-B4032E/2020|EPI_ISL_710877|2020 ···················· ···················· ···················· ···················· ···················· ····················
X 2306 hCoV-19/Wales/QEUH-B3F9A8/2020|EPI_ISL_710879|2020 ···················· ···················· ···················· ···················· ···················· ····················
X 2307 hCoV-19/Wales/QEUH-B3FDD9/2020|EPI_ISL_710880|2020 ···················· ···················· ···················· ···················· ···················· ····················
X 2308 hCoV-19/Wales/QEUH-B3F9D5/2020|EPI_ISL_710881|2020 ···················· ···················· ···················· ···················· ···················· ····················
X 2309 hCoV-19/Wales/QEUH-B3FD51/2020|EPI_ISL_710882|2020 ···················· ···················· ···················· ···················· ···················· ····················
X 2310 hCoV-19/Wales/QEUH-B3FA5A/2020|EPI_ISL_710883|2020 ···················· ···················· ···················· ···················· ···················· ····················
X 2311 hCoV-19/Wales/QEUH-B3F9B7/2020|EPI_ISL_710884|2020 ···················· ···················· ···················· ···················· ···················· ····················
X 2312 hCoV-19/Wales/QEUH-B3FD7F/2020|EPI_ISL_710885|2020 ···················· ···················· ···················· ···················· ···················· ····················
X 2313 hCoV-19/Wales/QEUH-B3FCBE/2020|EPI_ISL_710886|2020 ···················· ···················· ···················· ···················· ···················· ····················
X 2314 hCoV-19/Wales/ALDP-B4920A/2020|EPI_ISL_710887|2020 ···················· ···················· ···················· ···················· ···················· ····················
X 2315 hCoV-19/Wales/ALDP-B48845/2020|EPI_ISL_710900|2020 ···················· ···················· ···················· ···················· ···················· ····················
X 2316 hCoV-19/Wales/ALDP-B49273/2020|EPI_ISL_710901|2020 ···················· ···················· ···················· ···················· ···················· ····················
X 2317 hCoV-19/Wales/QEUH-B30B83/2020|EPI_ISL_710903|2020 ···················· ···················· ···················· ···················· ···················· ····················
X 2318 hCoV-19/Wales/QEUH-B2F9D8/2020|EPI_ISL_710904|2020 ···················· ···················· ···················· ···················· ···················· ····················
X 2319 hCoV-19/Wales/QEUH-B30AD1/2020|EPI_ISL_710905|2020 ···················· ···················· ···················· ···················· ···················· ····················
X 2320 hCoV-19/Wales/QEUH-B30B0B/2020|EPI_ISL_710906|2020 ···················· ···················· ···················· ···················· ···················· ····················
X 2321 hCoV-19/Wales/MILK-B3D193/2020|EPI_ISL_710907|2020 ···················· ···················· ···················· ···················· ···················· ····················
X 2322 hCoV-19/Wales/ALDP-B19FFD/2020|EPI_ISL_710908|2020 ···················· ···················· ···················· ···················· ···················· ····················
X 2323 hCoV-19/Wales/ALDP-B1A25D/2020|EPI_ISL_710909|2020 ···················· ···················· ···················· ···················· ···················· ····················
X 2324 hCoV-19/Wales/ALDP-B1B66F/2020|EPI_ISL_710910|2020 ···················· ···················· ···················· ···················· ···················· ····················
X 2325 hCoV-19/Wales/ALDP-B1A1E7/2020|EPI_ISL_710911|2020 ···················· ···················· ···················· ···················· ···················· ····················
X 2326 hCoV-19/Wales/ALDP-B19F84/2020|EPI_ISL_710912|2020 ···················· ···················· ···················· ···················· ···················· ····················
X 2327 hCoV-19/Wales/ALDP-B1B74E/2020|EPI_ISL_710913|2020 ···················· ···················· ···················· ···················· ···················· ····················

| | | | | | | | |
|---|---|---|---|---|---|---|---|
| X | 2328 hCoV-19/Wales/ALDP-B1B75D/2020\|EPI_ISL_710914\|2020 | ···················· | ···················· | ···················· | ···················· | ···················· | ···················· |
| X | 2329 hCoV-19/Wales/ALDP-B1A27B/2020\|EPI_ISL_710915\|2020 | ···················· | ···················· | ···················· | ···················· | ···················· | ···················· |
| X | 2330 hCoV-19/Wales/ALDP-B1A105/2020\|EPI_ISL_710916\|2020 | ···················· | ···················· | ···················· | ···················· | ···················· | ···················· |
| X | 2331 hCoV-19/Wales/ALDP-B1A6D9/2020\|EPI_ISL_710917\|2020 | ···················· | ···················· | ···················· | ···················· | ···················· | ···················· |
| X | 2332 hCoV-19/Wales/ALDP-B19F93/2020\|EPI_ISL_710918\|2020 | ···················· | ···················· | ···················· | ···················· | ···················· | ···················· |
| X | 2333 hCoV-19/Wales/ALDP-B1B122/2020\|EPI_ISL_710919\|2020 | ···················· | ···················· | ···················· | ···················· | ···················· | ···················· |
| X | 2334 hCoV-19/Wales/ALDP-B1E21D/2020\|EPI_ISL_710920\|2020 | ···················· | ···················· | ···················· | ···················· | ···················· | ···················· |
| X | 2335 hCoV-19/Wales/ALDP-B1A044/2020\|EPI_ISL_710921\|2020 | ···················· | ···················· | ···················· | ···················· | ···················· | ···················· |
| X | 2336 hCoV-19/Wales/ALDP-B19FC0/2020\|EPI_ISL_710922\|2020 | ···················· | ···················· | ···················· | ···················· | ···················· | ···················· |
| X | 2337 hCoV-19/Wales/ALDP-B1B6C9/2020\|EPI_ISL_710923\|2020 | ···················· | ···················· | ···················· | ···················· | ···················· | ···················· |
| X | 2338 hCoV-19/Wales/ALDP-B1A071/2020\|EPI_ISL_710924\|2020 | ···················· | ···················· | ···················· | ···················· | ···················· | ···················· |
| X | 2339 hCoV-19/Wales/ALDP-B1B6D8/2020\|EPI_ISL_710925\|2020 | ···················· | ···················· | ···················· | ···················· | ···················· | ···················· |
| X | 2340 hCoV-19/Wales/ALDP-B1A23F/2020\|EPI_ISL_710926\|2020 | ···················· | ···················· | ···················· | ···················· | ···················· | ···················· |
| X | 2341 hCoV-19/Wales/ALDP-B1A6E8/2020\|EPI_ISL_710927\|2020 | ···················· | ···················· | ···················· | ···················· | ···················· | ···················· |
| X | 2342 hCoV-19/Wales/ALDP-B1A24E/2020\|EPI_ISL_710928\|2020 | ···················· | ···················· | ···················· | ···················· | ···················· | ···················· |
| X | 2343 hCoV-19/Wales/ALDP-B1E1F2/2020\|EPI_ISL_710929\|2020 | ···················· | ···················· | ···················· | ···················· | ······T············· | ···················· |
| X | 2344 hCoV-19/Wales/ALDP-B1A18D/2020\|EPI_ISL_710930\|2020 | ···················· | ···················· | ···················· | ···················· | ···················· | ···················· |
| X | 2345 hCoV-19/Wales/ALDP-B1A1D8/2020\|EPI_ISL_710931\|2020 | ···················· | ···················· | ···················· | ···················· | ···················· | ···················· |
| X | 2346 hCoV-19/Wales/ALDP-B1B6F6/2020\|EPI_ISL_710932\|2020 | ···················· | ···················· | ···················· | ···················· | ···················· | ···················· |
| X | 2347 hCoV-19/Wales/ALDP-B1B3C2/2020\|EPI_ISL_710933\|2020 | ···················· | ···················· | ···················· | ···················· | ···················· | ···················· |
| X | 2348 hCoV-19/Wales/ALDP-B1B711/2020\|EPI_ISL_710934\|2020 | ···················· | ···················· | ···················· | ···················· | ···················· | ···················· |
| X | 2349 hCoV-19/Wales/ALDP-B1B6AB/2020\|EPI_ISL_710935\|2020 | ···················· | ···················· | ···················· | ···················· | ···················· | ···················· |
| X | 2350 hCoV-19/Wales/ALDP-B1B113/2020\|EPI_ISL_710936\|2020 | ···················· | ···················· | ···················· | ···················· | ···················· | ···················· |
| X | 2351 hCoV-19/Wales/ALDP-B1A6BB/2020\|EPI_ISL_710937\|2020 | ···················· | ···················· | ···················· | ···················· | ···················· | ···················· |
| X | 2352 hCoV-19/Wales/ALDP-B1A660/2020\|EPI_ISL_710938\|2020 | ···················· | ···················· | ···················· | ···················· | ···················· | ···················· |
| X | 2353 hCoV-19/Wales/ALDP-B1B641/2020\|EPI_ISL_710940\|2020 | ···················· | ···················· | ···················· | ···················· | ···················· | ···················· |
| X | 2354 hCoV-19/Wales/ALDP-B19FDF/2020\|EPI_ISL_710941\|2020 | ···················· | ···················· | ···················· | ···················· | ···················· | ···················· |
| X | 2355 hCoV-19/Wales/ALDP-B1E22C/2020\|EPI_ISL_710942\|2020 | ···················· | ···················· | ···················· | ···················· | ···················· | ···················· |
| X | 2356 hCoV-19/Wales/ALDP-B1B68D/2020\|EPI_ISL_710965\|2020 | ···················· | ···················· | ···················· | ···················· | ···················· | ···················· |
| X | 2357 hCoV-19/Wales/ALDP-B1B16E/2020\|EPI_ISL_710966\|2020 | ···················· | ···················· | ···················· | ···················· | ···················· | ···················· |
| X | 2358 hCoV-19/Wales/ALDP-B1E23B/2020\|EPI_ISL_710967\|2020 | ···················· | ···················· | ···················· | ···················· | ···················· | ···················· |
| X | 2359 hCoV-19/Wales/ALDP-B19FB1/2020\|EPI_ISL_710968\|2020 | ···················· | ···················· | ···················· | ···················· | ···················· | ···················· |
| X | 2360 hCoV-19/Wales/ALDP-B1E24A/2020\|EPI_ISL_710969\|2020 | ···················· | ···················· | ···················· | ···················· | ···················· | ···················· |
| X | 2361 hCoV-19/Wales/ALDP-B1A035/2020\|EPI_ISL_710970\|2020 | ···················· | ···················· | ···················· | ···················· | ···················· | ···················· |
| X | 2362 hCoV-19/Wales/ALDP-B1B76C/2020\|EPI_ISL_710971\|2020 | ···················· | ···················· | ···················· | ···················· | ···················· | ···················· |
| X | 2363 hCoV-19/Wales/ALDP-B19F66/2020\|EPI_ISL_710972\|2020 | ···················· | ···················· | ···················· | ···················· | ···················· | ···················· |
| X | 2364 hCoV-19/Wales/ALDP-B1E1A7/2020\|EPI_ISL_710973\|2020 | ···················· | ···················· | ···················· | ···················· | ···················· | ···················· |
| X | 2365 hCoV-19/Wales/ALDP-B1E259/2020\|EPI_ISL_710974\|2020 | ···················· | ···················· | ···················· | ···················· | ···················· | ···················· |
| X | 2366 hCoV-19/Wales/ALDP-B1B73F/2020\|EPI_ISL_710975\|2020 | ···················· | ···················· | ···················· | ···················· | ···················· | ···················· |
| X | 2367 hCoV-19/Denmark/DCGC-15909/2020\|EPI_ISL_714478\|202 | ···················· | ···················· | ···················· | ···················· | ···················· | ···················· |
| X | 2368 hCoV-19/Denmark/DCGC-15910/2020\|EPI_ISL_714479\|202 | ···················· | ···················· | ···················· | ···················· | ···················· | ···················· |
| X | 2369 hCoV-19/Denmark/DCGC-15911/2020\|EPI_ISL_714480\|202 | ···················· | ···················· | ···················· | ···················· | ···················· | ···················· |
| X | 2370 hCoV-19/Denmark/DCGC-15915/2020\|EPI_ISL_714481\|202 | ···················· | ···················· | ···················· | ···················· | ···················· | ···················· |
| X | 2371 hCoV-19/Denmark/DCGC-15918/2020\|EPI_ISL_714482\|202 | ···················· | ···················· | ···················· | ···················· | ···················· | ···················· |
| X | 2372 hCoV-19/Denmark/DCGC-15920/2020\|EPI_ISL_714483\|202 | ···················· | ···················· | ···················· | ···················· | ···················· | ···················· |
| X | 2373 hCoV-19/Denmark/DCGC-15921/2020\|EPI_ISL_714484\|202 | ···················· | ···················· | ···················· | ···················· | ······T············· | ···················· |
| X | 2374 hCoV-19/Denmark/DCGC-15922/2020\|EPI_ISL_714485\|202 | ···················· | ···················· | ···················· | ···················· | ···················· | ···················· |
| X | 2375 hCoV-19/Denmark/DCGC-15924/2020\|EPI_ISL_714486\|202 | ···················· | ···················· | ···················· | ···················· | ···················· | ···················· |
| X | 2376 hCoV-19/Denmark/DCGC-15926/2020\|EPI_ISL_714487\|202 | ···················· | ···················· | ···················· | ···················· | ···················· | ···················· |
| X | 2377 hCoV-19/Denmark/DCGC-15927/2020\|EPI_ISL_714488\|202 | ···················· | ···················· | ···················· | ···················· | ···················· | ···················· |
| X | 2378 hCoV-19/Denmark/DCGC-15929/2020\|EPI_ISL_714489\|202 | ···················· | ···················· | ···················· | ···················· | ···················· | ···················· |
| X | 2379 hCoV-19/Denmark/DCGC-15930/2020\|EPI_ISL_714490\|202 | ···················· | ···················· | ···················· | ···················· | ···················· | ···················· |
| X | 2380 hCoV-19/Denmark/DCGC-15933/2020\|EPI_ISL_714491\|202 | ···················· | ···················· | ···················· | ···················· | ···················· | ···················· |
| X | 2381 hCoV-19/Denmark/DCGC-15936/2020\|EPI_ISL_714492\|202 | ···················· | ···················· | ···················· | ···················· | ···················· | ···················· |
| X | 2382 hCoV-19/Denmark/DCGC-15938/2020\|EPI_ISL_714493\|202 | ···················· | ···················· | ···················· | ···················· | ···················· | ···················· |
| X | 2383 hCoV-19/Denmark/DCGC-15940/2020\|EPI_ISL_714494\|202 | ···················· | ···················· | ···················· | ···················· | ···················· | ···················· |
| X | 2384 hCoV-19/Denmark/DCGC-15942/2020\|EPI_ISL_714495\|202 | ···················· | ···················· | ···················· | ···················· | ···················· | ···················· |
| X | 2385 hCoV-19/Denmark/DCGC-15945/2020\|EPI_ISL_714496\|202 | ···················· | ···················· | ···················· | ···················· | ···················· | ···················· |
| X | 2386 hCoV-19/Denmark/DCGC-15946/2020\|EPI_ISL_714497\|202 | ···················· | ···················· | ···················· | ···················· | ···················· | ···················· |
| X | 2387 hCoV-19/Denmark/DCGC-15947/2020\|EPI_ISL_714498\|202 | ···················· | ···················· | ···················· | ···················· | ···················· | ···················· |
| X | 2388 hCoV-19/Denmark/DCGC-15948/2020\|EPI_ISL_714499\|202 | ···················· | ···················· | ···················· | ···················· | ···················· | ···················· |
| X | 2389 hCoV-19/England/CAMC-C424DD/2020\|EPI_ISL_719218\|20 | ···················· | ···················· | ···················· | ···················· | ···················· | ···················· |
| X | 2390 hCoV-19/England/CAMC-C44F07/2020\|EPI_ISL_719240\|20 | ···················· | ···················· | ···················· | ···················· | ···················· | ···················· |
| X | 2391 hCoV-19/England/CAMC-C43A9E/2020\|EPI_ISL_719245\|20 | ···················· | ···················· | ···················· | ···················· | ······T············· | ···················· |
| X | 2392 hCoV-19/England/CAMC-C3E6B0/2020\|EPI_ISL_719248\|20 | ···················· | ···················· | ···················· | ···················· | ···················· | ···················· |
| X | 2393 hCoV-19/England/CAMC-C3FA86/2020\|EPI_ISL_719249\|20 | ···················· | ···················· | ···················· | ···················· | ···················· | ···················· |
| X | 2394 hCoV-19/England/CAMC-C40E3B/2020\|EPI_ISL_719253\|20 | ···················· | ···················· | ···················· | ···················· | ···················· | ···················· |
| X | 2395 hCoV-19/England/CAMC-C3FAC2/2020\|EPI_ISL_719254\|20 | ···················· | ···················· | ···················· | ···················· | ···················· | ···················· |
| X | 2396 hCoV-19/England/CAMC-C3F0DF/2020\|EPI_ISL_719256\|20 | ···················· | ···················· | ···················· | ···················· | ···················· | ···················· |
| X | 2397 hCoV-19/England/CAMC-C43B22/2020\|EPI_ISL_719260\|20 | ···················· | ···················· | ···················· | ···················· | ···················· | ···················· |
| X | 2398 hCoV-19/England/CAMC-C4367C/2020\|EPI_ISL_719261\|20 | ···················· | ···················· | ···················· | ···················· | ······T············· | ···················· |
| X | 2399 hCoV-19/England/CAMC-C43B40/2020\|EPI_ISL_719275\|20 | ···················· | ···················· | ···················· | ···················· | ···················· | ···················· |
| X | 2400 hCoV-19/England/CAMC-C40B34/2020\|EPI_ISL_719276\|20 | ···················· | ···················· | ···················· | ···················· | ···················· | ···················· |
| X | 2401 hCoV-19/England/CAMC-C4422C/2020\|EPI_ISL_719281\|20 | ···················· | ···················· | ···················· | ···················· | ···················· | ···················· |
| X | 2402 hCoV-19/England/CAMC-C47A6D/2020\|EPI_ISL_719284\|20 | ···················· | ···················· | ···················· | ···················· | ···················· | ···················· |
| X | 2403 hCoV-19/England/CAMC-C43E56/2020\|EPI_ISL_719285\|20 | ···················· | ···················· | ···················· | ···················· | ···················· | ···················· |
| X | 2404 hCoV-19/England/CAMC-C3FBFC/2020\|EPI_ISL_719286\|20 | ···················· | ···················· | ···················· | ···················· | ···················· | ···················· |
| X | 2405 hCoV-19/England/CAMC-C3F3B8/2020\|EPI_ISL_719294\|20 | ···················· | ···················· | ···················· | ···················· | ···················· | ···················· |
| X | 2406 hCoV-19/England/CAMC-C45258/2020\|EPI_ISL_719300\|20 | ···················· | ···················· | ···················· | ···················· | ···················· | ···················· |
| X | 2407 hCoV-19/England/CAMC-C3FC35/2020\|EPI_ISL_719301\|20 | ···················· | ···················· | ···················· | ···················· | ···················· | ···················· |

| | | | | | | | | |
|---|---|---|---|---|---|---|---|---|
| X | 2408 | hCoV-19/England/CAMC-C40132/2020\|EPI_ISL_719305\|20 | ···················· | ····A··············· | ····················· | ···················· | ···················· | ···················· |
| X | 2409 | hCoV-19/England/CAMC-C3F3D6/2020\|EPI_ISL_719307\|20 | ···················· | ···················· | ····················· | ···················· | ···················· | ···················· |
| X | 2410 | hCoV-19/England/CAMC-C4430B/2020\|EPI_ISL_719308\|20 | ···················· | ···················· | ····················· | ···················· | ···················· | ···················· |
| X | 2411 | hCoV-19/England/MILK-BB16F2/2020\|EPI_ISL_720200\|20 | ···················· | ···················· | ····················· | ···················· | ···················· | ···················· |
| X | 2412 | hCoV-19/England/MILK-BB2758/2020\|EPI_ISL_720201\|20 | ···················· | ···················· | ····················· | ···················· | ···················· | ···················· |
| X | 2413 | hCoV-19/England/MILK-BB207B/2020\|EPI_ISL_720202\|20 | ···················· | ···················· | ····················· | ···················· | ···················· | ···················· |
| X | 2414 | hCoV-19/England/MILK-BB0435/2020\|EPI_ISL_720204\|20 | ···················· | ···················· | ····················· | ···················· | ···················· | ···················· |
| X | 2415 | hCoV-19/England/MILK-BB1F51/2020\|EPI_ISL_720230\|20 | ···················· | ···················· | ····················· | ···················· | ···················· | ···················· |
| X | 2416 | hCoV-19/England/MILK-BB180B/2020\|EPI_ISL_720233\|20 | ···················· | ···················· | ····················· | ···················· | ···················· | ···················· |
| X | 2417 | hCoV-19/England/MILK-BB204E/2020\|EPI_ISL_720234\|20 | ···················· | ···················· | ····················· | ···················· | ···················· | ···················· |
| X | 2418 | hCoV-19/England/MILK-BB12B2/2020\|EPI_ISL_720235\|20 | ···················· | ···················· | ····················· | ···················· | ·······T············ | ···················· |
| X | 2419 | hCoV-19/England/MILK-BB159B/2020\|EPI_ISL_720236\|20 | ···················· | ···················· | ····················· | ···················· | ···················· | ···················· |
| X | 2420 | hCoV-19/England/MILK-BB04AE/2020\|EPI_ISL_720238\|20 | ···················· | ···················· | ····················· | ···················· | ···················· | ···················· |
| X | 2421 | hCoV-19/England/MILK-BB1E90/2020\|EPI_ISL_720239\|20 | ···················· | ···················· | ····················· | ···················· | ···················· | ···················· |
| X | 2422 | hCoV-19/England/MILK-BB18FC/2020\|EPI_ISL_720240\|20 | ···················· | ···················· | ····················· | ···················· | ···················· | ···················· |
| X | 2423 | hCoV-19/England/MILK-BB0FAD/2020\|EPI_ISL_720241\|20 | ···················· | ···················· | ····················· | ···················· | ···················· | ···················· |
| X | 2424 | hCoV-19/England/MILK-BB1F9D/2020\|EPI_ISL_720242\|20 | ···················· | ···················· | ····················· | ···················· | ···················· | ···················· |
| X | 2425 | hCoV-19/England/MILK-BB0F70/2020\|EPI_ISL_720243\|20 | ···················· | ···················· | ····················· | ···················· | ···················· | ···················· |
| X | 2426 | hCoV-19/England/MILK-BB014D/2020\|EPI_ISL_720244\|20 | ···················· | ···················· | ····················· | ···················· | ·······T············ | ···················· |
| X | 2427 | hCoV-19/England/MILK-BB1407/2020\|EPI_ISL_720247\|20 | ···················· | ···················· | ····················· | ···················· | ···················· | ···················· |
| X | 2428 | hCoV-19/England/MILK-BB14DA/2020\|EPI_ISL_720248\|20 | ···················· | ···················· | ····················· | ···················· | ···················· | ···················· |
| X | 2429 | hCoV-19/England/MILK-BB1030/2020\|EPI_ISL_720251\|20 | ···················· | ···················· | ····················· | ···················· | ···················· | ···················· |
| X | 2430 | hCoV-19/England/MILK-BB012F/2020\|EPI_ISL_720253\|20 | ···················· | ···················· | ····················· | ···················· | ···················· | ···················· |
| X | 2431 | hCoV-19/England/MILK-BB2530/2020\|EPI_ISL_720254\|20 | ···················· | ···················· | ····················· | ···················· | ···················· | ···················· |
| X | 2432 | hCoV-19/England/MILK-BB15AA/2020\|EPI_ISL_720255\|20 | ···················· | ···················· | ····················· | ···················· | ·······T············ | ···················· |
| X | 2433 | hCoV-19/England/MILK-BB17C2/2020\|EPI_ISL_720256\|20 | ···················· | ·A·················· | ····················· | ···················· | ···················· | ···A················ |
| X | 2434 | hCoV-19/England/MILK-BB17B3/2020\|EPI_ISL_720258\|20 | ···················· | ···················· | ····················· | ···················· | ···················· | ···················· |
| X | 2435 | hCoV-19/England/MILK-BB1294/2020\|EPI_ISL_720262\|20 | ·········T·········· | ···················· | ····················· | ···················· | ···················· | ···················· |
| X | 2436 | hCoV-19/England/MILK-BB18ED/2020\|EPI_ISL_720263\|20 | ···················· | ···················· | ····················· | ···················· | ···················· | ···················· |
| X | 2437 | hCoV-19/England/MILK-BB20C6/2020\|EPI_ISL_720264\|20 | ···················· | ···················· | ····················· | ···················· | ···················· | ···················· |
| X | 2438 | hCoV-19/England/MILK-BB261F/2020\|EPI_ISL_720266\|20 | ···················· | ···················· | ····················· | ···················· | ···················· | ···················· |
| X | 2439 | hCoV-19/England/MILK-BB1ECD/2020\|EPI_ISL_720274\|20 | ···················· | ···················· | ····················· | ···················· | ···················· | ···················· |
| X | 2440 | hCoV-19/England/MILK-BB12A3/2020\|EPI_ISL_720275\|20 | ···················· | ···················· | ····················· | ···················· | ···················· | ···················· |
| X | 2441 | hCoV-19/England/MILK-BB1883/2020\|EPI_ISL_720276\|20 | ···················· | ···················· | ····················· | ···················· | ···················· | ···················· |
| X | 2442 | hCoV-19/England/MILK-BB1513/2020\|EPI_ISL_720277\|20 | ···················· | ···················· | ····················· | ···················· | ···················· | ···················· |
| X | 2443 | hCoV-19/England/MILK-BB03FC/2020\|EPI_ISL_720278\|20 | ···················· | ···················· | ····················· | ···················· | ···················· | ···················· |
| X | 2444 | hCoV-19/England/MILK-BB1416/2020\|EPI_ISL_720279\|20 | ···················· | ···················· | ····················· | ···················· | ···················· | ···················· |
| X | 2445 | hCoV-19/England/MILK-BB2776/2020\|EPI_ISL_720285\|20 | ···················· | ···················· | ····················· | ···················· | ···················· | ···················· |
| X | 2446 | hCoV-19/England/MILK-BB15B9/2020\|EPI_ISL_720286\|20 | ···················· | ···················· | ····················· | ···················· | ···················· | ···················· |
| X | 2447 | hCoV-19/England/MILK-BB1003/2020\|EPI_ISL_720289\|20 | ···················· | ···················· | ····················· | ···················· | ···················· | ···················· |
| X | 2448 | hCoV-19/England/MILK-BB0408/2020\|EPI_ISL_720295\|20 | ···················· | ···················· | ····················· | ···················· | ···················· | ···················· |
| X | 2449 | hCoV-19/England/MILK-BB12EF/2020\|EPI_ISL_720296\|20 | ···················· | ···················· | ····················· | ···················· | ···················· | ···················· |
| X | 2450 | hCoV-19/England/MILK-BB2460/2020\|EPI_ISL_720297\|20 | ···················· | ···················· | ····················· | ···················· | ···················· | ···················· |
| X | 2451 | hCoV-19/England/MILK-BB259A/2020\|EPI_ISL_720298\|20 | ···················· | ···················· | ····················· | ···················· | ···················· | ···················· |
| X | 2452 | hCoV-19/England/ALDP-BC8397/2020\|EPI_ISL_721514\|20 | ···················· | ···················· | ····················· | ···················· | ···················· | ···················· |
| X | 2453 | hCoV-19/England/ALDP-BC850A/2020\|EPI_ISL_721515\|20 | ···················· | ···················· | ····················· | ···················· | ···················· | ···················· |
| X | 2454 | hCoV-19/England/ALDP-C06138/2020\|EPI_ISL_721517\|20 | ···················· | ···················· | ····················· | ···················· | ···················· | ···················· |
| X | 2455 | hCoV-19/England/QEUH-C17C9D/2020\|EPI_ISL_721519\|20 | ···················· | ···················· | ····················· | ···················· | ···················· | ···················· |
| X | 2456 | hCoV-19/England/ALDP-BE9381/2020\|EPI_ISL_721525\|20 | ···················· | ···················· | ····················· | ···················· | ···················· | ···················· |
| X | 2457 | hCoV-19/England/MILK-BEE39B/2020\|EPI_ISL_721526\|20 | ···················· | ···················· | ····················· | ···················· | ···················· | ···················· |
| X | 2458 | hCoV-19/England/MILK-BE6EDE/2020\|EPI_ISL_721529\|20 | ···················· | ···················· | ····················· | ···················· | ···················· | ···················· |
| X | 2459 | hCoV-19/England/QEUH-BC35C3/2020\|EPI_ISL_721533\|20 | ···················· | ····A··············· | ····················· | ···················· | ···················· | ···················· |
| X | 2460 | hCoV-19/England/QEUH-B2BC88/2020\|EPI_ISL_721534\|20 | ···················· | ···················· | ····················· | ···················· | ···················· | ···················· |
| X | 2461 | hCoV-19/England/QEUH-B0F6F5/2020\|EPI_ISL_721535\|20 | ···················· | ···················· | ····················· | ···················· | ···················· | ···················· |
| X | 2462 | hCoV-19/England/QEUH-A9EBEE/2020\|EPI_ISL_721558\|20 | ···················· | ···················· | ····················· | ···················· | ···················· | ···················· |
| X | 2463 | hCoV-19/England/QEUH-A9F28E/2020\|EPI_ISL_721559\|20 | ···················· | ···················· | ····················· | ···················· | ···················· | ···················· |
| X | 2464 | hCoV-19/England/QEUH-A9ED8E/2020\|EPI_ISL_721560\|20 | ···················· | ···················· | ····················· | ···················· | ···················· | ···················· |
| X | 2465 | hCoV-19/England/QEUH-A9EB57/2020\|EPI_ISL_721561\|20 | ···················· | ···················· | ····················· | ···················· | ···················· | ···················· |
| X | 2466 | hCoV-19/England/QEUH-A9EB1B/2020\|EPI_ISL_721563\|20 | ···················· | ···················· | ····················· | ···················· | ···················· | ···················· |
| X | 2467 | hCoV-19/England/QEUH-A9EAD2/2020\|EPI_ISL_721564\|20 | ···················· | ···················· | ····················· | ···················· | ···················· | ···················· |
| X | 2468 | hCoV-19/England/QEUH-A9EAE1/2020\|EPI_ISL_721565\|20 | ···················· | ···················· | ····················· | ···················· | ···················· | ···················· |
| X | 2469 | hCoV-19/England/QEUH-A9F0B1/2020\|EPI_ISL_721566\|20 | ···················· | ···················· | ····················· | ···················· | ···················· | ···················· |
| X | 2470 | hCoV-19/England/QEUH-A9F27F/2020\|EPI_ISL_721567\|20 | ···················· | ···················· | ····················· | ···················· | ···················· | ···················· |
| X | 2471 | hCoV-19/Japan/IC-0353/2020\|EPI_ISL_721568\|2020-12 | ···················· | ···················· | ····················· | ···················· | ···················· | ···················· |
| X | 2472 | hCoV-19/Japan/IC-0361/2020\|EPI_ISL_721569\|2020-11 | ···················· | ···················· | ····················· | ···················· | ···················· | ···················· |
| X | 2473 | hCoV-19/Japan/IC-0355/2020\|EPI_ISL_721575\|2020-12 | ···················· | ···················· | ····················· | ···················· | ···················· | ···················· |
| X | 2474 | hCoV-19/Japan/IC-0329/2020\|EPI_ISL_721577\|2020-11 | ···················· | ···················· | ····················· | ···················· | ···················· | ···················· |
| X | 2475 | hCoV-19/Japan/IC-0331/2020\|EPI_ISL_721578\|2020-11 | ···················· | ···················· | ····················· | ···················· | ···················· | ···················· |
| X | 2476 | hCoV-19/Japan/IC-0332/2020\|EPI_ISL_721579\|2020-11 | ···················· | ···················· | ····················· | ···················· | ···················· | ···················· |
| X | 2477 | hCoV-19/Japan/IC-0333/2020\|EPI_ISL_721580\|2020-11 | ···················· | ···················· | ····················· | ···················· | ···················· | ···················· |
| X | 2478 | hCoV-19/Japan/IC-0334/2020\|EPI_ISL_721581\|2020-11 | ···················· | ···················· | ····················· | ···················· | ···················· | ···················· |
| X | 2479 | hCoV-19/Japan/IC-0335/2020\|EPI_ISL_721582\|2020-11 | ···················· | ···················· | ····················· | ···················· | ···················· | ···················· |
| X | 2480 | hCoV-19/Japan/IC-0336/2020\|EPI_ISL_721583\|2020-11 | ···················· | ···················· | ····················· | ···················· | ···················· | ···················· |
| X | 2481 | hCoV-19/Japan/IC-0337/2020\|EPI_ISL_721584\|2020-11 | ···················· | ·A·················· | ····················· | ···················· | ···················· | ···A················ |
| X | 2482 | hCoV-19/Japan/IC-0338/2020\|EPI_ISL_721585\|2020-11 | ···················· | ·A·················· | ····················· | ···················· | ···················· | ···A················ |
| X | 2483 | hCoV-19/Japan/IC-0339/2020\|EPI_ISL_721586\|2020-11 | ···················· | ···················· | ····················· | ···················· | ···················· | ···················· |
| X | 2484 | hCoV-19/Japan/IC-0342/2020\|EPI_ISL_721587\|2020-11 | ···················· | ·A·················· | ····················· | ···················· | ···················· | ···A················ |
| X | 2485 | hCoV-19/Japan/IC-0343/2020\|EPI_ISL_721588\|2020-11 | ···················· | ···················· | ····················· | ···················· | ···················· | ···················· |
| X | 2486 | hCoV-19/Japan/IC-0344/2020\|EPI_ISL_721589\|2020-11 | ···················· | ···················· | ····················· | ···················· | ···················· | ···················· |
| X | 2487 | hCoV-19/Japan/IC-0345/2020\|EPI_ISL_721590\|2020-11 | ···················· | ···················· | ····················· | ···················· | ···················· | ···················· |

| | | | | | | | |
|---|---|---|---|---|---|---|---|
| X | 2488 hCoV-19/Japan/IC-0346/2020\|EPI_ISL_721591\|2020-11 | ···················· | ·A·················· | ···················· | ···················· | ···················· | ···A················ |
| X | 2489 hCoV-19/Japan/IC-0347/2020\|EPI_ISL_721592\|2020-11 | ···················· | ···················· | ···················· | ···················· | ···················· | ···················· |
| X | 2490 hCoV-19/Japan/IC-0348/2020\|EPI_ISL_721593\|2020-11 | ···················· | ···················· | ···················· | ···················· | ···················· | ···················· |
| X | 2491 hCoV-19/Japan/IC-0349/2020\|EPI_ISL_721594\|2020-11 | ···················· | ·A·················· | ···················· | ···················· | ···················· | ···A················ |
| X | 2492 hCoV-19/Japan/IC-0350/2020\|EPI_ISL_721595\|2020-11 | ···················· | ···················· | ···················· | ···················· | ···················· | ···················· |
| X | 2493 hCoV-19/Japan/IC-0351/2020\|EPI_ISL_721596\|2020-11 | ···················· | ···················· | ···················· | ···················· | ···················· | ···················· |
| X | 2494 hCoV-19/Japan/IC-0352/2020\|EPI_ISL_721597\|2020-11 | ···················· | ···················· | ···················· | ···················· | ···················· | ···················· |
| X | 2495 hCoV-19/Japan/IC-0354/2020\|EPI_ISL_721598\|2020-12 | ···················· | ···················· | ···················· | ···················· | ···················· | ···················· |
| X | 2496 hCoV-19/Japan/IC-0356/2020\|EPI_ISL_721599\|2020-12 | ···················· | ·A··A··············· | ···················· | ···················· | ···················· | ···················· |
| X | 2497 hCoV-19/Japan/IC-0377/2020\|EPI_ISL_721617\|2020-12 | ···················· | ···················· | ···················· | ···················· | ···················· | ···················· |
| X | 2498 hCoV-19/Japan/IC-0378/2020\|EPI_ISL_721618\|2020-11 | ···················· | ···················· | ···················· | ···················· | ···················· | ···················· |
| X | 2499 hCoV-19/Japan/IC-0379/2020\|EPI_ISL_721619\|2020-11 | ···················· | ···················· | ···················· | ···················· | ···················· | ···················· |
| X | 2500 hCoV-19/Japan/IC-0380/2020\|EPI_ISL_721620\|2020-11 | ···················· | ···················· | ···················· | ···················· | ···················· | ···················· |
| X | 2501 hCoV-19/Japan/IC-0382/2020\|EPI_ISL_721621\|2020-11 | ···················· | ···················· | ···················· | ···················· | ···················· | ···················· |
| X | 2502 hCoV-19/Japan/IC-0383/2020\|EPI_ISL_721622\|2020-12 | ···················· | ·A·················· | ···················· | ···················· | ···················· | ···················· |
| X | 2503 hCoV-19/Japan/IC-0384/2020\|EPI_ISL_721623\|2020-12 | ···················· | ···················· | ···················· | ···················· | ···················· | ···················· |
| X | 2504 hCoV-19/Netherlands/OV-EMC-99/2020\|EPI_ISL_722845\| | ···················· | ···················· | ···················· | ···················· | ·····T·············· | ···················· |
| X | 2505 hCoV-19/Netherlands/ZH-EMC-1039/2020\|EPI_ISL_72284 | ···················· | ···················· | ···················· | ···················· | ···················· | ···················· |
| X | 2506 hCoV-19/Netherlands/OV-EMC-94/2020\|EPI_ISL_722847\| | ···················· | ···················· | ···················· | ···················· | ···················· | ···················· |
| X | 2507 hCoV-19/Netherlands/GE-EMC-432/2020\|EPI_ISL_722848 | ···················· | ···················· | ···················· | ···················· | ···················· | ···················· |
| X | 2508 hCoV-19/Netherlands/GE-EMC-433/2020\|EPI_ISL_722849 | ···················· | ···················· | ···················· | ···················· | ···················· | ···················· |
| X | 2509 hCoV-19/Italy/APU-IZSPB-153APT/2020\|EPI_ISL_722856 | ···················· | ···················· | ···················· | ···················· | ···················· | ···················· |
| X | 2510 hCoV-19/Italy/APU-IZSPB-156APT/2020\|EPI_ISL_722857 | ···················· | ···················· | ···················· | ···················· | ···················· | ···················· |
| X | 2511 hCoV-19/Italy/APU-IZSPB-159APT/2020\|EPI_ISL_722858 | ···················· | ···················· | ···················· | ···················· | ···················· | ···················· |
| X | 2512 hCoV-19/Italy/APU-IZSPB-178PT/2020\|EPI_ISL_722859\| | ···················· | ···················· | ···················· | ···················· | ···················· | ···················· |
| X | 2513 hCoV-19/Italy/APU-IZSPB-181PT/2020\|EPI_ISL_722860\| | ···················· | ···················· | ···················· | ···················· | ···················· | ···················· |
| X | 2514 hCoV-19/Italy/APU-IZSPB-183PT/2020\|EPI_ISL_722861\| | ···················· | ···················· | ···················· | ···················· | ···················· | ···················· |
| X | 2515 hCoV-19/Italy/APU-IZSPB-184PT/2020\|EPI_ISL_722862\| | ···················· | ···················· | ···················· | ···················· | ···················· | ···················· |
| X | 2516 hCoV-19/Italy/APU-IZSPB-185PT/2020\|EPI_ISL_722863\| | ···················· | ···················· | ···················· | ···················· | ···················· | ···················· |
| X | 2517 hCoV-19/Italy/APU-IZSPB-186PT/2020\|EPI_ISL_722864\| | ···················· | ···················· | ···················· | ···················· | ···················· | ···················· |
| X | 2518 hCoV-19/Italy/APU-IZSPB-188PT/2020\|EPI_ISL_722865\| | ···················· | ···················· | ···················· | ···················· | ···················· | ···················· |
| X | 2519 hCoV-19/Italy/APU-IZSPB-189PT/2020\|EPI_ISL_722866\| | ···················· | ···················· | ···················· | ···················· | ···················· | ···················· |
| X | 2520 hCoV-19/Italy/BAS-IZSPB-390PT/2020\|EPI_ISL_722889\| | ···················· | ···················· | ···················· | ···················· | ···················· | ···················· |
| X | 2521 hCoV-19/Italy/BAS-IZSPB-391PT/2020\|EPI_ISL_722890\| | ···················· | ···················· | ···················· | ···················· | ·····T·············· | ···················· |
| X | 2522 hCoV-19/Italy/BAS-IZSPB-398PT/2020\|EPI_ISL_722891\| | ···················· | ···················· | ···················· | ···················· | ···················· | ···················· |
| X | 2523 hCoV-19/Italy/BAS-IZSPB-194PT/2020\|EPI_ISL_722892\| | ···················· | ···················· | ···················· | ···················· | ···················· | ···················· |
| X | 2524 hCoV-19/Italy/APU-IZSPB-332PT/2020\|EPI_ISL_722893\| | ···················· | ···················· | ···················· | ···················· | ···················· | ···················· |
| X | 2525 hCoV-19/Italy/APU-IZSPB-333PT/2020\|EPI_ISL_722894\| | ···················· | ···················· | ···················· | ···················· | ···················· | ···················· |
| X | 2526 hCoV-19/Italy/APU-IZSPB-187PT/2020\|EPI_ISL_722895\| | ···················· | ···················· | ···················· | ···················· | ···················· | ···················· |
| X | 2527 hCoV-19/Italy/APU-IZSPB-142APT/2020\|EPI_ISL_722896 | ···················· | ···················· | ···················· | ···················· | ···················· | ···················· |
| X | 2528 hCoV-19/Italy/APU-IZSPB-151APT/2020\|EPI_ISL_722897 | ···················· | ···················· | ···················· | ···················· | ···················· | ···················· |
| X | 2529 hCoV-19/Italy/APU-IZSPB-180PT/2020\|EPI_ISL_722898\| | ···················· | ···················· | ···················· | ···················· | ···················· | ···················· |
| X | 2530 hCoV-19/Italy/APU-IZSPB-150PT/2020\|EPI_ISL_722899\| | ···················· | ···················· | ···················· | ···················· | ···················· | ···················· |
| X | 2531 hCoV-19/Italy/APU-IZSPB-196PT/2020\|EPI_ISL_722900\| | ···················· | ···················· | ···················· | ···················· | ···················· | ···················· |
| X | 2532 hCoV-19/Italy/BAS-IZSPB-336PT/2020\|EPI_ISL_722901\| | ···················· | ···················· | ···················· | ···················· | ···················· | ···················· |
| X | 2533 hCoV-19/Italy/BAS-IZSPB-357PT/2020\|EPI_ISL_722902\| | ···················· | ·A·················· | ···················· | ···················· | ···················· | ···················· |
| X | 2534 hCoV-19/Italy/BAS-IZSPB-358PT/2020\|EPI_ISL_722903\| | ···················· | ···················· | ···················· | ···················· | ···················· | ···················· |
| X | 2535 hCoV-19/Italy/BAS-IZSPB-370PT/2020\|EPI_ISL_722904\| | ···················· | ···················· | ···················· | ···················· | ···················· | ···················· |
| X | 2536 hCoV-19/Italy/BAS-IZSPB-372PT/2020\|EPI_ISL_722905\| | ···················· | ···················· | ···················· | ···················· | ···················· | ···················· |
| X | 2537 hCoV-19/Italy/BAS-IZSPB-379PT/2020\|EPI_ISL_722906\| | ···················· | ···················· | ···················· | ···················· | ···················· | ···················· |
| X | 2538 hCoV-19/Italy/BAS-IZSPB-382PT/2020\|EPI_ISL_722907\| | ···················· | ···················· | ···················· | ···················· | ···················· | ···················· |
| X | 2539 hCoV-19/Italy/APU-IZSPB-330PT/2020\|EPI_ISL_722908\| | ···················· | ···················· | ···················· | ···················· | ···················· | ···················· |
| X | 2540 hCoV-19/Italy/APU-IZSPB-334PT/2020\|EPI_ISL_722909\| | ···················· | ···················· | ···················· | ···················· | ···················· | ···················· |
| X | 2541 hCoV-19/Italy/APU-IZSPB-335PT/2020\|EPI_ISL_722910\| | ···················· | ···················· | ···················· | ···················· | ···················· | ···················· |
| X | 2542 hCoV-19/Italy/APU-IZSPB-331PT/2020\|EPI_ISL_722911\| | ···················· | ···················· | ···················· | ···················· | ···················· | ···················· |
| X | 2543 hCoV-19/Italy/APU-IZSPB-149APT/2020\|EPI_ISL_722912 | ···················· | ···················· | ···················· | ···················· | ···················· | ···················· |
| X | 2544 hCoV-19/Italy/BAS-IZSPB-352PT/2020\|EPI_ISL_722913\| | ···················· | ···················· | ···················· | ···················· | ···················· | ···················· |
| X | 2545 hCoV-19/Italy/BAS-IZSPB-356PT/2020\|EPI_ISL_722914\| | ···················· | ···················· | ···················· | ···················· | ···················· | ···················· |
| X | 2546 hCoV-19/Italy/BAS-IZSPB-365PT/2020\|EPI_ISL_722915\| | ···················· | ···················· | ···················· | ···················· | ···················· | ···················· |
| X | 2547 hCoV-19/Italy/BAS-IZSPB-366PT/2020\|EPI_ISL_722916\| | ···················· | ···················· | ···················· | ···················· | ···················· | ···················· |
| X | 2548 hCoV-19/Italy/BAS-IZSPB-371PT/2020\|EPI_ISL_722917\| | ···················· | ···················· | ···················· | ···················· | ···················· | ···················· |
| X | 2549 hCoV-19/Italy/APU-IZSPB-377PT/2020\|EPI_ISL_722918\| | ···················· | ···················· | ···················· | ···················· | ·····T·············· | ···················· |
| X | 2550 hCoV-19/Italy/BAS-IZSPB-378PT/2020\|EPI_ISL_722919\| | ···················· | ···················· | ···················· | ···················· | ···················· | ···················· |
| X | 2551 hCoV-19/Italy/BAS-IZSPB-386PT/2020\|EPI_ISL_722920\| | ···················· | ···················· | ···················· | ···················· | ···················· | ···················· |
| X | 2552 hCoV-19/Italy/BAS-IZSPB-387PT/2020\|EPI_ISL_722921\| | ···················· | ···················· | ···················· | ···················· | ···················· | ···················· |
| X | 2553 hCoV-19/Belgium/ULG-11027/2020\|EPI_ISL_722945\|2020 | ···················· | ···················· | ···················· | ···················· | ···················· | ···················· |
| X | 2554 hCoV-19/Belgium/ULG-11031/2020\|EPI_ISL_722948\|2020 | ···················· | ···················· | ···················· | ···················· | ·····T·············· | ···················· |
| X | 2555 hCoV-19/Belgium/ULG-11032/2020\|EPI_ISL_722949\|2020 | ···················· | ···················· | ···················· | ···················· | ·····T·············· | ···················· |
| X | 2556 hCoV-19/Belgium/ULG-11033/2020\|EPI_ISL_722950\|2020 | ···················· | ···················· | ···················· | ···················· | ···················· | ···················· |
| X | 2557 hCoV-19/Belgium/ULG-11037/2020\|EPI_ISL_722951\|2020 | ···················· | ···················· | ···················· | ···················· | ···················· | ···················· |
| X | 2558 hCoV-19/Belgium/ULG-11038/2020\|EPI_ISL_722952\|2020 | ···················· | ···················· | ···················· | ···················· | ···················· | ···················· |
| X | 2559 hCoV-19/Belgium/ULG-11039/2020\|EPI_ISL_722953\|2020 | ···················· | ···················· | ···················· | ···················· | ···C················ | ···················· |
| X | 2560 hCoV-19/Belgium/ULG-11040/2020\|EPI_ISL_722954\|2020 | ···················· | ···················· | ···················· | ···················· | ···················· | ···················· |
| X | 2561 hCoV-19/Wales/ALDP-B1B67E/2020\|EPI_ISL_710943\|2020 | ···················· | ···················· | ···················· | ···················· | ···················· | ···················· |
| X | 2562 hCoV-19/Wales/ALDP-B1E295/2020\|EPI_ISL_710944\|2020 | ···················· | ···················· | ···················· | ···················· | ···················· | ···················· |
| X | 2563 hCoV-19/Wales/ALDP-B1E268/2020\|EPI_ISL_710945\|2020 | ···················· | ···················· | ···················· | ···················· | ···················· | ···················· |
| X | 2564 hCoV-19/Wales/ALDP-B1B104/2020\|EPI_ISL_710946\|2020 | ···················· | ···················· | ···················· | ···················· | ···················· | ···················· |
| X | 2565 hCoV-19/Wales/ALDP-B1B69C/2020\|EPI_ISL_710947\|2020 | ···················· | ···················· | ···················· | ···················· | ···················· | ···················· |
| X | 2566 hCoV-19/Wales/ALDP-B1A220/2020\|EPI_ISL_710948\|2020 | ···················· | ···················· | ···················· | ···················· | ···················· | ···················· |
| X | 2567 hCoV-19/Wales/ALDP-B1E2D1/2020\|EPI_ISL_710949\|2020 | ···················· | ···················· | ···················· | ···················· | ···················· | ···················· |

| | | | | | | | | |
|---|---|---|---|---|---|---|---|---|
| X | 2568 | hCoV-19/Wales/ALDP-B1B702/2020\|EPI_ISL_710950\|2020 | ···················· | ···················· | ···················· | ···················· | ···················· | ···················· |
| X | 2569 | hCoV-19/Wales/ALDP-B1A017/2020\|EPI_ISL_710951\|2020 | ···················· | ···················· | ···················· | ···················· | ···················· | ···················· |
| X | 2570 | hCoV-19/Wales/ALDP-B1E2A4/2020\|EPI_ISL_710952\|2020 | ···················· | ···················· | ···················· | ···················· | ···················· | ···················· |
| X | 2571 | hCoV-19/Wales/ALDP-B19F1B/2020\|EPI_ISL_710953\|2020 | ···················· | ···················· | ···················· | ···················· | ···················· | ···················· |
| X | 2572 | hCoV-19/Wales/ALDP-B1E1E3/2020\|EPI_ISL_710954\|2020 | ···················· | ···················· | ···················· | ···················· | ···················· | ···················· |
| X | 2573 | hCoV-19/Wales/ALDP-B1A68E/2020\|EPI_ISL_710955\|2020 | ···················· | ···················· | ···················· | ···················· | ···················· | ···················· |
| X | 2574 | hCoV-19/Wales/ALDP-B1E286/2020\|EPI_ISL_710956\|2020 | ···················· | ···················· | ···················· | ···················· | ···················· | ···················· |
| X | 2575 | hCoV-19/Wales/ALDP-B1B0F8/2020\|EPI_ISL_710958\|2020 | ···················· | ···················· | ···················· | ···················· | ···················· | ···················· |
| X | 2576 | hCoV-19/Wales/ALDP-B1E2C2/2020\|EPI_ISL_710959\|2020 | ···················· | ···················· | ···················· | ···················· | ···················· | ···················· |
| X | 2577 | hCoV-19/Wales/ALDP-B1B720/2020\|EPI_ISL_710960\|2020 | ···················· | ···················· | ···················· | ···················· | ···················· | ···················· |
| X | 2578 | hCoV-19/Wales/ALDP-B1E1B6/2020\|EPI_ISL_710961\|2020 | ···················· | ···················· | ···················· | ···················· | ···················· | ···················· |
| X | 2579 | hCoV-19/Wales/ALDP-B1A211/2020\|EPI_ISL_710963\|2020 | ···················· | ···················· | ···················· | ···················· | ···················· | ···················· |
| X | 2580 | hCoV-19/Wales/ALDP-B1B632/2020\|EPI_ISL_710964\|2020 | ···················· | ···················· | ···················· | ···················· | ···················· | ···················· |
| X | 2581 | hCoV-19/Wales/ALDP-B1A008/2020\|EPI_ISL_710977\|2020 | ···················· | ···················· | ···················· | ···················· | ···················· | ···················· |
| X | 2582 | hCoV-19/Wales/ALDP-B1B650/2020\|EPI_ISL_710979\|2020 | ···················· | ···················· | ···················· | ···················· | ···················· | ···················· |
| X | 2583 | hCoV-19/Wales/ALDP-B1A6CA/2020\|EPI_ISL_710980\|2020 | ···················· | ···················· | ···················· | ···················· | ···················· | ···················· |
| X | 2584 | hCoV-19/Wales/ALDP-B1A26C/2020\|EPI_ISL_710981\|2020 | ···················· | ···················· | ···················· | ···················· | ···················· | ···················· |
| X | 2585 | hCoV-19/Wales/ALDP-B1D58E/2020\|EPI_ISL_710982\|2020 | ···················· | ···················· | ···················· | ···················· | ···················· | ···················· |
| X | 2586 | hCoV-19/Wales/ALDP-B1D5D9/2020\|EPI_ISL_710983\|2020 | ···················· | ···················· | ···················· | ···················· | ···················· | ···················· |
| X | 2587 | hCoV-19/Wales/ALDP-B1D5E8/2020\|EPI_ISL_710984\|2020 | ···················· | ···················· | ···················· | ···················· | ···················· | ···················· |
| X | 2588 | hCoV-19/Wales/ALDP-B1FA9C/2020\|EPI_ISL_710985\|2020 | ···················· | ···················· | ···················· | ···················· | ···················· | ···················· |
| X | 2589 | hCoV-19/Wales/ALDP-B1D621/2020\|EPI_ISL_710986\|2020 | ···················· | ···················· | ···················· | ···················· | ···················· | ···················· |
| X | 2590 | hCoV-19/Wales/ALDP-B1F58C/2020\|EPI_ISL_710987\|2020 | ···················· | ···················· | ···················· | ···················· | ···················· | ···················· |
| X | 2591 | hCoV-19/Wales/ALDP-B1D57F/2020\|EPI_ISL_710988\|2020 | ···················· | ···················· | ···················· | ···················· | ···················· | ···················· |
| X | 2592 | hCoV-19/Wales/ALDP-B1F865/2020\|EPI_ISL_710990\|2020 | ···················· | ···················· | ···················· | ···················· | ···················· | ···················· |
| X | 2593 | hCoV-19/Wales/ALDP-B1BC5E/2020\|EPI_ISL_710991\|2020 | ···················· | ···················· | ···················· | ···················· | ···················· | ···················· |
| X | 2594 | hCoV-19/Wales/ALDP-B1D59D/2020\|EPI_ISL_710992\|2020 | ···················· | ···················· | ···················· | ···················· | ···················· | ···················· |
| X | 2595 | hCoV-19/Wales/ALDP-B1D524/2020\|EPI_ISL_710993\|2020 | ···················· | ···················· | ···················· | ···················· | ···················· | ···················· |
| X | 2596 | hCoV-19/Wales/QEUH-B2E12C/2020\|EPI_ISL_710995\|2020 | ···················· | ···················· | ···················· | ···················· | ···················· | ···················· |
| X | 2597 | hCoV-19/Wales/QEUH-B2E0A7/2020\|EPI_ISL_710996\|2020 | ···················· | ···················· | ···················· | ···················· | ···················· | ···················· |
| X | 2598 | hCoV-19/Wales/QEUH-B2E1D1/2020\|EPI_ISL_710997\|2020 | ···················· | ···················· | ···················· | ···················· | ···················· | ···················· |
| X | 2599 | hCoV-19/Wales/QEUH-B2E1FF/2020\|EPI_ISL_710998\|2020 | ···················· | ···················· | ···················· | ···················· | ···················· | ···················· |
| X | 2600 | hCoV-19/Wales/QEUH-B2E4AB/2020\|EPI_ISL_710999\|2020 | ···················· | ···················· | ···················· | ···················· | ···················· | ···················· |
| X | 2601 | hCoV-19/Belgium/UGent-261/2020\|EPI_ISL_718000\|2020 | ···················· | ···················· | ···················· | ···················· | ···················· | ···················· |
| X | 2602 | hCoV-19/Belgium/UGent-263/2020\|EPI_ISL_718001\|2020 | ···················· | ···················· | ···················· | ···················· | ···················· | ···················· |
| X | 2603 | hCoV-19/Belgium/UGent-264/2020\|EPI_ISL_718002\|2020 | ···················· | ···················· | ···················· | ···················· | ···················· | ···················· |
| X | 2604 | hCoV-19/Belgium/UGent-265/2020\|EPI_ISL_718003\|2020 | ···················· | ···················· | ···················· | ···················· | ···················· | ···················· |
| X | 2605 | hCoV-19/Belgium/UGent-266/2020\|EPI_ISL_718004\|2020 | ···················· | ···················· | ···················· | ···················· | ···················· | ···················· |
| X | 2607 | hCoV-19/Belgium/UGent-268/2020\|EPI_ISL_718006\|2020 | ···················· | ···················· | ···················· | ···················· | ···················· | ···················· |
| X | 2608 | hCoV-19/Belgium/UGent-269/2020\|EPI_ISL_718007\|2020 | ···················· | ···················· | ···················· | ···················· | ···················· | ···················· |
| X | 2610 | hCoV-19/Belgium/UGent-272/2020\|EPI_ISL_718009\|2020 | ···················· | ···················· | ···················· | ···················· | ···················· | ···················· |
| X | 2612 | hCoV-19/Belgium/UGent-275/2020\|EPI_ISL_718011\|2020 | ···················· | ···················· | ···················· | ···················· | ···················· | ···················· |
| X | 2613 | hCoV-19/Belgium/UGent-276/2020\|EPI_ISL_718012\|2020 | ···················· | ···················· | ···················· | ···················· | ···················· | ···················· |
| X | 2615 | hCoV-19/Belgium/UGent-280/2020\|EPI_ISL_718014\|2020 | ···················· | ···················· | ···················· | ···················· | ···················· | ···················· |
| X | 2616 | hCoV-19/Belgium/UGent-281/2020\|EPI_ISL_718015\|2020 | ···················· | ···················· | ···················· | ···················· | ···················· | ···················· |
| X | 2617 | hCoV-19/Belgium/UGent-286/2020\|EPI_ISL_718016\|2020 | ···················· | ···················· | ···················· | ···················· | ···················· | ···················· |
| X | 2618 | hCoV-19/Belgium/UGent-287/2020\|EPI_ISL_718017\|2020 | ···················· | ···················· | ···················· | ···················· | ···················· | ···················· |
| X | 2619 | hCoV-19/Belgium/UGent-288/2020\|EPI_ISL_718018\|2020 | ···················· | ···················· | ···················· | ···················· | ···················· | ···················· |
| X | 2621 | hCoV-19/Belgium/UGent-291/2020\|EPI_ISL_718020\|2020 | ···················· | ···················· | ···················· | ···················· | ···················· | ···················· |
| X | 2622 | hCoV-19/Belgium/UGent-292/2020\|EPI_ISL_718021\|2020 | ···················· | ···················· | ···················· | ···················· | ···················· | ···················· |
| X | 2624 | hCoV-19/Belgium/UGent-295/2020\|EPI_ISL_718023\|2020 | ···················· | ···················· | ···················· | ···················· | ···················· | ···················· |
| X | 2625 | hCoV-19/Belgium/UGent-297/2020\|EPI_ISL_718024\|2020 | ···················· | ···················· | ···················· | ···················· | ···················· | ···················· |
| X | 2626 | hCoV-19/Belgium/UGent-299/2020\|EPI_ISL_718025\|2020 | ···················· | ···················· | ···················· | ···················· | ···················· | ···················· |
| X | 2627 | hCoV-19/Belgium/UGent-302/2020\|EPI_ISL_718026\|2020 | ···················· | ···················· | ···················· | ···················· | ···················· | ···················· |
| X | 2628 | hCoV-19/Germany/NW-HHU-331/2020\|EPI_ISL_718028\|202 | ···················· | ···················· | ···················· | ···················· | ···················· | ···················· |
| X | 2629 | hCoV-19/Germany/NW-HHU-337/2020\|EPI_ISL_718029\|202 | ···················· | ···················· | ···················· | ···················· | ···················· | ···················· |
| X | 2630 | hCoV-19/Germany/NW-HHU-328/2020\|EPI_ISL_718030\|202 | ···················· | ···················· | ···················· | ···················· | ···················· | ···················· |
| X | 2631 | hCoV-19/Germany/NW-HHU-332/2020\|EPI_ISL_718033\|202 | ···················· | ···················· | ···················· | ···················· | ···················· | ···················· |
| X | 2632 | hCoV-19/Germany/NW-HHU-333/2020\|EPI_ISL_718034\|202 | ···················· | ···················· | ···················· | ···················· | ···················· | ···················· |
| X | 2633 | hCoV-19/Germany/NW-HHU-335/2020\|EPI_ISL_718035\|202 | ···················· | ···················· | ···················· | ···················· | ···················· | ···················· |
| X | 2634 | hCoV-19/Germany/NW-HHU-336/2020\|EPI_ISL_718036\|202 | ···················· | ···················· | ···················· | ···················· | ···················· | ···················· |
| X | 2635 | hCoV-19/Germany/NW-HHU-338/2020\|EPI_ISL_718037\|202 | ···················· | ···················· | ···················· | ···················· | ···················· | ···················· |
| X | 2636 | hCoV-19/Germany/NW-HHU-339/2020\|EPI_ISL_718038\|202 | ···················· | ···················· | ···················· | ···················· | ···················· | ···················· |
| X | 2637 | hCoV-19/Germany/NW-HHU-341/2020\|EPI_ISL_718039\|202 | ···················· | ···················· | ···················· | ···················· | ···················· | ···················· |
| X | 2638 | hCoV-19/Germany/NW-HHU-373/2020\|EPI_ISL_718056\|202 | ···················· | ···················· | ···················· | ···················· | ···················· | ···················· |
| X | 2639 | hCoV-19/Germany/NW-HHU-374/2020\|EPI_ISL_718057\|202 | ···················· | ···················· | ···················· | ···················· | ···················· | ···················· |
| X | 2640 | hCoV-19/Germany/NW-HHU-382/2020\|EPI_ISL_718058\|202 | ···················· | ···················· | ···················· | ···················· | ···················· | ···················· |
| X | 2641 | hCoV-19/Germany/NW-HHU-385/2020\|EPI_ISL_718059\|202 | ···················· | ···················· | ···················· | ···················· | ···················· | ···················· |
| X | 2642 | hCoV-19/Germany/NW-HHU-407/2020\|EPI_ISL_718062\|202 | ···················· | ···················· | ···················· | ···················· | ···················· | ···················· |
| X | 2643 | hCoV-19/Germany/NW-HHU-408/2020\|EPI_ISL_718063\|202 | ···················· | ···················· | ···················· | ···················· | ···················· | ···················· |
| X | 2644 | hCoV-19/Germany/NW-HHU-386/2020\|EPI_ISL_718068\|202 | ···················· | ···················· | ···················· | ···················· | ···················· | ···················· |
| X | 2645 | hCoV-19/Germany/NW-HHU-387/2020\|EPI_ISL_718069\|202 | ···················· | ···················· | ···················· | ···················· | ···················· | ···················· |
| X | 2646 | hCoV-19/England/CAMC-C3F321/2020\|EPI_ISL_719315\|20 | ···················· | ···················· | ···················· | ···················· | ···················· | ···················· |
| X | 2647 | hCoV-19/England/CAMC-C3FC9F/2020\|EPI_ISL_719317\|20 | ···················· | ···················· | ···················· | ···················· | ···················· | ···················· |
| X | 2648 | hCoV-19/England/CAMC-C3F312/2020\|EPI_ISL_719322\|20 | ···················· | ···················· | ···················· | ···················· | ···················· | ···················· |
| X | 2649 | hCoV-19/England/CAMC-C448DF/2020\|EPI_ISL_719326\|20 | ···················· | ···················· | ···················· | ···················· | ···················· | ···················· |
| X | 2650 | hCoV-19/England/CAMC-C43E65/2020\|EPI_ISL_719330\|20 | ···················· | ···················· | ···················· | ···················· | ······T············· | ···················· |
| X | 2651 | hCoV-19/England/CAMC-C456E3/2020\|EPI_ISL_719333\|20 | ···················· | ···················· | ···················· | ···················· | ···················· | ···················· |
| X | 2652 | hCoV-19/England/CAMC-C3F60A/2020\|EPI_ISL_719334\|20 | ···················· | ···················· | ···················· | ···················· | ···················· | ···················· |
| X | 2653 | hCoV-19/England/CAMC-C43E92/2020\|EPI_ISL_719343\|20 | ···················· | ···················· | ···················· | ···················· | ···················· | ···················· |

```
X 2654 hCoV-19/England/CAMC-C43EDE/2020|EPI_ISL_719344|20 .................... .................... .................... .................... .................... ....................
X 2655 hCoV-19/England/CAMC-C3FEF3/2020|EPI_ISL_719348|20 .................... .A.................. .................... .................... .................... ...A................
X 2656 hCoV-19/England/CAMC-C43F62/2020|EPI_ISL_719353|20 .................... .A.................. .................... .................... .................... ....................
X 2657 hCoV-19/England/CAMC-C4130E/2020|EPI_ISL_719356|20 .................... .................... .................... .................... ......T............. ....................
X 2658 hCoV-19/England/CAMC-C3FC53/2020|EPI_ISL_719360|20 .................... .................... .................... .................... .................... ....................
X 2659 hCoV-19/England/CAMC-C43EB0/2020|EPI_ISL_719366|20 .................... .................... .................... .................... .................... ....................
X 2660 hCoV-19/England/CAMC-C3FB92/2020|EPI_ISL_719368|20 .................... .................... .................... .................... .................... ....................
X 2661 hCoV-19/England/CAMC-C4574A/2020|EPI_ISL_719380|20 .................... .................... .................... .................... .................... ....................
X 2662 hCoV-19/England/CAMC-C3F5D0/2020|EPI_ISL_719381|20 .................... .................... .................... .................... .................... ....................
X 2663 hCoV-19/England/CAMC-C4125C/2020|EPI_ISL_719383|20 .................... .................... .................... .................... .................... ....................
X 2664 hCoV-19/England/CAMC-C4325A/2020|EPI_ISL_719387|20 .................... .................... .................... .................... .................... ....................
X 2665 hCoV-19/England/CAMC-C43278/2020|EPI_ISL_719388|20 .................... .................... .................... .................... .................... ....................
X 2666 hCoV-19/England/CAMC-C3FC71/2020|EPI_ISL_719389|20 .................... .................... .................... .................... .................... ....................
X 2667 hCoV-19/England/CAMC-C455AA/2020|EPI_ISL_719395|20 .................... .................... .................... .................... .................... ....................
X 2668 hCoV-19/England/CAMC-C40105/2020|EPI_ISL_719396|20 .................... .................... .................... .................... .................... ....................
X 2669 hCoV-19/England/CAMC-C44839/2020|EPI_ISL_719398|20 .................... .A.................. .................... .................... .................... ...A................
X 2670 hCoV-19/England/CAMC-C3F5FE/2020|EPI_ISL_719399|20 .................... .................... .................... .................... .................... ....................
X 2671 hCoV-19/England/MILK-BB15C8/2020|EPI_ISL_720300|20 .................... .................... .................... .................... .................... ....................
X 2672 hCoV-19/England/MILK-BB1531/2020|EPI_ISL_720305|20 .................... .................... .................... .................... .................... ....................
X 2673 hCoV-19/England/MILK-BB19AE/2020|EPI_ISL_720306|20 .................... .................... .................... .................... .................... ....................
X 2674 hCoV-19/England/MILK-BB1E18/2020|EPI_ISL_720308|20 .................... .................... .................... .................... .................... ....................
X 2675 hCoV-19/England/MILK-BB262E/2020|EPI_ISL_720309|20 .................... .................... .................... .................... .................... ....................
X 2676 hCoV-19/England/MILK-BB2442/2020|EPI_ISL_720314|20 .................... .................... .................... .................... .................... ....................
X 2677 hCoV-19/England/MILK-BB1012/2020|EPI_ISL_720315|20 .................... .................... .................... .................... .................... ....................
X 2678 hCoV-19/England/MILK-BB2512/2020|EPI_ISL_720316|20 .................... .................... .................... .................... .................... ....................
X 2679 hCoV-19/England/MILK-BB2600/2020|EPI_ISL_720317|20 .................... .................... .................... .................... .................... ....................
X 2680 hCoV-19/England/MILK-BB1847/2020|EPI_ISL_720320|20 .................... .................... .................... .................... .................... ....................
X 2681 hCoV-19/England/MILK-BB2415/2020|EPI_ISL_720321|20 .................... .................... .................... .................... .................... ....................
X 2682 hCoV-19/England/MILK-BB04F9/2020|EPI_ISL_720322|20 .................... .................... .................... .................... .................... ....................
X 2683 hCoV-19/England/MILK-BB0909/2020|EPI_ISL_720323|20 .................... .A.................. .................... .................... .................... ...A................
X 2684 hCoV-19/England/MILK-BB1F33/2020|EPI_ISL_720324|20 .................... .................... .................... .................... .................... ....................
X 2685 hCoV-19/England/MILK-BB265B/2020|EPI_ISL_720325|20 .................... .................... .................... .................... .................... ....................
X 2686 hCoV-19/England/MILK-BB08B1/2020|EPI_ISL_720327|20 .................... .................... .................... .................... .................... ....................
X 2687 hCoV-19/England/MILK-BB1829/2020|EPI_ISL_720330|20 .................... .................... .................... .................... .................... ....................
X 2688 hCoV-19/England/MILK-BB0990/2020|EPI_ISL_720332|20 .................... .................... .................... .................... .................... ....................
X 2689 hCoV-19/England/MILK-BAC2E7/2020|EPI_ISL_720334|20 .................... .................... .................... .................... .................... ....................
X 2690 hCoV-19/England/MILK-BB221B/2020|EPI_ISL_720336|20 .................... .................... .................... .................... .................... ....................
X 2691 hCoV-19/England/MILK-BB03B0/2020|EPI_ISL_720337|20 .................... .................... .................... .................... .................... ....................
X 2692 hCoV-19/England/MILK-BB0E64/2020|EPI_ISL_720338|20 .................... .................... .................... .................... .................... ....................
X 2693 hCoV-19/England/MILK-BAFCB9/2020|EPI_ISL_720339|20 .................... .................... .................... .................... .................... ....................
X 2694 hCoV-19/England/MILK-BB10E5/2020|EPI_ISL_720340|20 .................... .................... .................... .................... .................... ....................
X 2695 hCoV-19/England/MILK-BB213C/2020|EPI_ISL_720341|20 .................... .................... .................... .................... ......T............. ....................
X 2696 hCoV-19/England/MILK-BB0EA0/2020|EPI_ISL_720342|20 .................... .................... .................... .................... .................... ....................
X 2697 hCoV-19/England/MILK-BB2327/2020|EPI_ISL_720343|20 .................... .................... .................... .................... .................... ....................
X 2698 hCoV-19/England/MILK-BAC8B8/2020|EPI_ISL_720344|20 .................... .................... .................... .................... .................... ....................
X 2699 hCoV-19/England/MILK-BB0365/2020|EPI_ISL_720347|20 .................... .................... .................... .................... ......T............. ....................
X 2700 hCoV-19/England/MILK-BB0B30/2020|EPI_ISL_720348|20 .................... .................... .................... .................... .................... ....................
X 2701 hCoV-19/England/MILK-BB1D48/2020|EPI_ISL_720349|20 .................... .................... .................... .................... .................... ....................
X 2702 hCoV-19/England/MILK-BAFAA0/2020|EPI_ISL_720350|20 .................... .................... .................... .................... .................... ....................
X 2703 hCoV-19/England/MILK-BAC88B/2020|EPI_ISL_720352|20 .................... .................... .................... .................... .................... ....................
X 2704 hCoV-19/England/MILK-BB0CA6/2020|EPI_ISL_720353|20 .................... .................... .................... .................... .................... ....................
X 2705 hCoV-19/England/MILK-BB1DDF/2020|EPI_ISL_720355|20 .................... .................... .................... .................... .................... ....................
X 2706 hCoV-19/England/MILK-BB06C6/2020|EPI_ISL_720356|20 .................... .................... .................... .................... .................... ....................
X 2707 hCoV-19/England/MILK-BB0866/2020|EPI_ISL_720357|20 .................... .................... .................... .................... .................... ....................
X 2708 hCoV-19/England/MILK-BAFC31/2020|EPI_ISL_720358|20 .................... .A.................. .................... .................... .................... ....................
X 2709 hCoV-19/England/MILK-BB0927/2020|EPI_ISL_720371|20 .................... .................... .................... .................... .................... ....................
X 2710 hCoV-19/England/MILK-BAFCC8/2020|EPI_ISL_720372|20 .................... .................... .................... .................... .................... ....................
X 2711 hCoV-19/England/MILK-BAFADD/2020|EPI_ISL_720373|20 .................... .................... .................... .................... .................... ....................
X 2712 hCoV-19/England/MILK-BAB831/2020|EPI_ISL_720374|20 .................... .................... .................... .................... .................... ....................
X 2713 hCoV-19/England/MILK-BB0BF4/2020|EPI_ISL_720375|20 .................... .................... .................... .................... ......T............. ....................
X 2714 hCoV-19/England/MILK-BAC51B/2020|EPI_ISL_720376|20 .................... .................... .................... .................... .................... ....................
X 2715 hCoV-19/England/MILK-BB0857/2020|EPI_ISL_720377|20 .................... .................... .................... .................... .................... ....................
X 2716 hCoV-19/England/MILK-BB2363/2020|EPI_ISL_720378|20 .................... .................... .................... .................... .................... ....................
X 2717 hCoV-19/England/MILK-BB0E19/2020|EPI_ISL_720379|20 .................... .................... .................... .................... .................... ....................
X 2718 hCoV-19/England/MILK-BABF2D/2020|EPI_ISL_720380|20 .................... .................... .................... .................... .................... ....................
X 2719 hCoV-19/England/MILK-BB0374/2020|EPI_ISL_720381|20 .................... .A.................. .................... .................... .................... ....................
X 2720 hCoV-19/England/MILK-BB1B3F/2020|EPI_ISL_720383|20 .................... .................... .................... .................... .................... ....................
X 2721 hCoV-19/England/MILK-BAC86D/2020|EPI_ISL_720384|20 .................... .................... .................... .................... .................... ....................
X 2722 hCoV-19/England/MILK-BABA0E/2020|EPI_ISL_720386|20 .................... .................... .................... .................... .................... ....................
X 2723 hCoV-19/England/MILK-BB10F4/2020|EPI_ISL_720387|20 .................... .................... .................... .................... .................... ....................
X 2724 hCoV-19/England/MILK-BAC3F3/2020|EPI_ISL_720389|20 .................... .................... .................... .................... .................... ....................
X 2725 hCoV-19/Japan/IC-0357/2020|EPI_ISL_721600|2020-12 .................... .................... .................... .................... .................... ....................
X 2726 hCoV-19/Japan/IC-0358/2020|EPI_ISL_721601|2020-11 .................... .................... .................... .................... .................... ....................
X 2727 hCoV-19/Japan/IC-0359/2020|EPI_ISL_721602|2020-11 .................... .................... .................... .................... .................... ....................
X 2728 hCoV-19/Japan/IC-0360/2020|EPI_ISL_721603|2020-11 .................... .................... .................... .................... .................... ....................
X 2729 hCoV-19/Japan/IC-0362/2020|EPI_ISL_721604|2020-11 .................... .................... .................... .................... .................... ....................
X 2730 hCoV-19/Japan/IC-0363/2020|EPI_ISL_721605|2020-11 .................... .................... .................... .................... .................... ....................
X 2731 hCoV-19/Japan/IC-0364/2020|EPI_ISL_721606|2020-11 .................... .................... .................... .................... .................... ....................
X 2732 hCoV-19/Japan/IC-0365/2020|EPI_ISL_721607|2020-11 .................... .................... .................... .................... .................... ....................
X 2733 hCoV-19/Japan/IC-0366/2020|EPI_ISL_721608|2020-11 .................... .................... .................... .................... .................... ....................
```

| | | | | | | | |
|---|---|---|---|---|---|---|---|
| X | 2734 hCoV-19/Japan/IC-0367/2020\|EPI_ISL_721609\|2020-11 | .................... | .................... | .................... | .................... | .................... | .................... |
| X | 2735 hCoV-19/Japan/IC-0368/2020\|EPI_ISL_721610\|2020-11 | .................... | .................... | .................... | .................... | .................... | .................... |
| X | 2736 hCoV-19/Japan/IC-0370/2020\|EPI_ISL_721611\|2020-12 | .................... | .................... | .................... | .................... | .................... | .................... |
| X | 2737 hCoV-19/Japan/IC-0371/2020\|EPI_ISL_721612\|2020-12 | .................... | .................... | .................... | .................... | .................... | .................... |
| X | 2738 hCoV-19/Japan/IC-0372/2020\|EPI_ISL_721613\|2020-12 | .................... | .A.................. | .................... | .................... | .................... | ...A................ |
| X | 2739 hCoV-19/Japan/IC-0373/2020\|EPI_ISL_721614\|2020-12 | .................... | .................... | .................... | .................... | .................... | .................... |
| X | 2740 hCoV-19/Japan/IC-0375/2020\|EPI_ISL_721615\|2020-12 | .................... | .............A...... | .................... | .................... | .................... | .................... |
| X | 2741 hCoV-19/Japan/IC-0376/2020\|EPI_ISL_721616\|2020-12 | .................... | .A.................. | .................... | .................... | .................... | .................... |
| X | 2743 hCoV-19/Italy/LAZ-INMI-102/2020\|EPI_ISL_721625\|202 | .................... | .................... | .................... | .................... | .................... | .................... |
| X | 2744 hCoV-19/Italy/LAZ-INMI-103/2020\|EPI_ISL_721626\|202 | .................... | .................... | .................... | .................... | ......T............. | .................... |
| X | 2745 hCoV-19/Italy/LAZ-INMI-104/2020\|EPI_ISL_721627\|202 | .................... | .................... | .................... | .................... | .................... | .................... |
| X | 2747 hCoV-19/India/MH-AFCO-14752/2020\|EPI_ISL_721629\|20 | .................... | .................... | .................... | .................... | .................... | .................... |
| X | 2748 hCoV-19/India/MH-AFCO-14778/2020\|EPI_ISL_721630\|20 | .................... | .................... | .................... | .................... | .................... | .................... |
| X | 2749 hCoV-19/India/MH-AFCO-15230/2020\|EPI_ISL_721631\|20 | .................... | .................... | .................... | .................... | .................... | .................... |
| X | 2750 hCoV-19/India/MH-AFCO-15275/2020\|EPI_ISL_721632\|20 | .................... | .................... | .................... | .................... | .................... | .................... |
| X | 2751 hCoV-19/India/MH-AFCO-15381/2020\|EPI_ISL_721633\|20 | .................... | .................... | .................... | .................... | .................... | .................... |
| X | 2752 hCoV-19/India/MH-AFCO-15524/2020\|EPI_ISL_721634\|20 | .................... | .................... | .................... | .................... | .................... | .................... |
| X | 2753 hCoV-19/India/MH-AFCO-15531/2020\|EPI_ISL_721635\|20 | .................... | .................... | .................... | .................... | .................... | .................... |
| X | 2754 hCoV-19/India/MH-AFCO-15533/2020\|EPI_ISL_721636\|20 | .................... | .................... | .................... | .................... | .................... | .................... |
| X | 2755 hCoV-19/India/MH-AFCO-8108/2020\|EPI_ISL_721637\|202 | .................... | .................... | .................... | .................... | .................... | .................... |
| X | 2756 hCoV-19/India/MH-AFCO-8189/2020\|EPI_ISL_721638\|202 | .................... | .................... | .................... | .................... | .................... | .................... |
| X | 2757 hCoV-19/India/MH-AFCO-8677/2020\|EPI_ISL_721639\|202 | ....T............... | .................... | .................... | .................... | .................... | .................... |
| X | 2758 hCoV-19/India/MH-AFCO-9102/2020\|EPI_ISL_721640\|202 | .................... | .................... | .................... | .................... | .................... | .................... |
| X | 2759 hCoV-19/India/MH-AFCO-9104/2020\|EPI_ISL_721641\|202 | .................... | .................... | .................... | .................... | .................... | .................... |
| X | 2760 hCoV-19/India/MH-AFCO-9315/2020\|EPI_ISL_721642\|202 | .................... | .................... | .................... | .................... | .................... | .................... |
| X | 2761 hCoV-19/India/MH-AFCO-9320/2020\|EPI_ISL_721643\|202 | .................... | .................... | .................... | .................... | .................... | .................... |
| X | 2762 hCoV-19/India/MH-AFCO-9325/2020\|EPI_ISL_721644\|202 | .................... | .................... | .................... | .................... | .................... | .................... |
| X | 2763 hCoV-19/India/MH-AFCO-9995/2020\|EPI_ISL_721645\|202 | .................... | .................... | .................... | .................... | .................... | .................... |
| X | 2764 hCoV-19/India/MH-NCCS-NR-10609/2020\|EPI_ISL_721646 | .................... | .................... | .................... | .................... | .................... | .................... |
| X | 2765 hCoV-19/India/MH-NCCS-NR-10951/2020\|EPI_ISL_721647 | .................... | .A.................. | .................... | .................... | .................... | .................... |
| X | 2766 hCoV-19/India/MH-NCCS-NR-11448/2020\|EPI_ISL_721648 | .................... | .................... | .................... | .................... | .................... | .................... |
| X | 2767 hCoV-19/India/MH-NCCS-NR-11688/2020\|EPI_ISL_721649 | .................... | .................... | .................... | .................... | .................... | .................... |
| X | 2768 hCoV-19/India/MH-NCCS-NR-11843/2020\|EPI_ISL_721650 | .................... | .................... | .................... | .................... | .................... | .................... |
| X | 2769 hCoV-19/India/MH-NCCS-NR-12620/2020\|EPI_ISL_721651 | .................... | .................... | .................... | .................... | .................... | .................... |
| X | 2770 hCoV-19/India/MH-NCCS-NR-13749/2020\|EPI_ISL_721653 | .................... | .................... | .................... | .................... | .................... | .................... |
| X | 2771 hCoV-19/India/MH-NCCS-NR-14003/2020\|EPI_ISL_721654 | .................... | .................... | .................... | .................... | .................... | .................... |
| X | 2772 hCoV-19/India/MH-NCCS-NR-14607/2020\|EPI_ISL_721655 | .................... | .................... | .................... | .................... | .................... | .................... |
| X | 2773 hCoV-19/India/MH-NCCS-NR-15788/2020\|EPI_ISL_721656 | .................... | .................... | .................... | .................... | .................... | .................... |
| X | 2774 hCoV-19/India/MH-NCCS-NR-4614/2020\|EPI_ISL_721657\| | .................... | .................... | .................... | .................... | .................... | .................... |
| X | 2775 hCoV-19/India/MH-NCCS-NR-6668/2020\|EPI_ISL_721659\| | .................... | .................... | .................... | .................... | .................... | .................... |
| X | 2776 hCoV-19/India/MH-NCCS-NR-7123/2020\|EPI_ISL_721660\| | .................... | .................... | .................... | .................... | .................... | .................... |
| X | 2777 hCoV-19/India/MH-NCCS-NR-7614/2020\|EPI_ISL_721661\| | .................... | .................... | .................... | .................... | .................... | .................... |
| X | 2778 hCoV-19/India/MH-NCCS-NR-7631/2020\|EPI_ISL_721662\| | .................... | .................... | .................... | .................... | .................... | .................... |
| X | 2779 hCoV-19/India/MH-NCCS-NR-7636/2020\|EPI_ISL_721663\| | .................... | .................... | .................... | .................... | .................... | .................... |
| X | 2780 hCoV-19/India/MH-NCCS-NR-8720/2020\|EPI_ISL_721664\| | .................... | .................... | .................... | .................... | .................... | .................... |
| X | 2781 hCoV-19/India/MH-NCCS-NR-8737/2020\|EPI_ISL_721665\| | .................... | .................... | .................... | .................... | .................... | .................... |
| X | 2782 hCoV-19/India/MH-NCCS-NR-9077/2020\|EPI_ISL_721666\| | .................... | .................... | .................... | .................... | .................... | .................... |
| X | 2783 hCoV-19/India/MH-NCCS-NR-9234/2020\|EPI_ISL_721667\| | .................... | .................... | .................... | .................... | .................... | .................... |
| X | 2784 hCoV-19/Switzerland/AG-ETHZ-340141/2020\|EPI_ISL_72 | .................... | .................... | .................... | .................... | .................... | .................... |
| X | 2785 hCoV-19/Switzerland/SO-ETHZ-380288/2020\|EPI_ISL_72 | .................... | .................... | .................... | .................... | .................... | .................... |
| X | 2786 hCoV-19/Switzerland/BL-ETHZ-340136/2020\|EPI_ISL_72 | .................... | .................... | .................... | .................... | .................... | .................... |
| X | 2787 hCoV-19/Switzerland/SO-ETHZ-390227/2020\|EPI_ISL_72 | .................... | .................... | .................... | .................... | .................... | .................... |
| X | 2788 hCoV-19/Italy/BAS-IZSPB-392PT/2020\|EPI_ISL_722922\| | .................... | .................... | .................... | .................... | .................... | .................... |
| X | 2789 hCoV-19/Italy/BAS-IZSPB-393PT/2020\|EPI_ISL_722923\| | .................... | .................... | .................... | .................... | .................... | .................... |
| X | 2790 hCoV-19/Italy/BAS-IZSPB-395PT/2020\|EPI_ISL_722924\| | .................... | .................... | .................... | .................... | .................... | .................... |
| X | 2791 hCoV-19/Italy/BAS-IZSPB-397PT/2020\|EPI_ISL_722925\| | .................... | .................... | .................... | .................... | .................... | .................... |
| X | 2792 hCoV-19/Belgium/ULG-11016/2020\|EPI_ISL_722934\|2020 | .................... | .................... | .................... | .................... | .................... | .................... |
| X | 2793 hCoV-19/Belgium/ULG-11029/2020\|EPI_ISL_722935\|2020 | .................... | .................... | .................... | .................... | .................... | .................... |
| X | 2794 hCoV-19/Belgium/ULG-11017/2020\|EPI_ISL_722936\|2020 | .................... | .................... | .................... | .................... | .................... | .................... |
| X | 2795 hCoV-19/Belgium/ULG-11018/2020\|EPI_ISL_722937\|2020 | .................... | .................... | .................... | .................... | .................... | .................... |
| X | 2796 hCoV-19/Belgium/ULG-11020/2020\|EPI_ISL_722938\|2020 | .................... | .................... | .................... | .................... | .................... | .................... |
| X | 2797 hCoV-19/Belgium/ULG-11021/2020\|EPI_ISL_722939\|2020 | .................... | .................... | .................... | .................... | .................... | .................... |
| X | 2798 hCoV-19/Belgium/ULG-11022/2020\|EPI_ISL_722940\|2020 | .................... | .................... | .................... | .................... | .................... | .................... |
| X | 2799 hCoV-19/Belgium/ULG-11023/2020\|EPI_ISL_722941\|2020 | .................... | .................... | .................... | .................... | .................... | .................... |
| X | 2800 hCoV-19/Belgium/ULG-11024/2020\|EPI_ISL_722942\|2020 | .................... | .................... | .................... | .................... | .................... | .................... |
| X | 2801 hCoV-19/Belgium/ULG-11025/2020\|EPI_ISL_722943\|2020 | .................... | .................... | .................... | .................... | ......T............. | .................... |
| X | 2802 hCoV-19/Belgium/ULG-11026/2020\|EPI_ISL_722944\|2020 | .................... | .................... | .................... | .................... | .................... | .................... |
| X | 2803 hCoV-19/Belgium/ULG-11028/2020\|EPI_ISL_722946\|2020 | .................... | .................... | .................... | .................... | .................... | .................... |
| X | 2804 hCoV-19/Belgium/ULG-11030/2020\|EPI_ISL_722947\|2020 | .................... | .................... | .................... | .................... | .................... | .................... |
| X | 2805 hCoV-19/Belgium/ULG-11042/2020\|EPI_ISL_722955\|2020 | .................... | .................... | .................... | .................... | ......T............. | .................... |
| X | 2806 hCoV-19/Belgium/ULG-11043/2020\|EPI_ISL_722956\|2020 | .................... | .................... | .................... | .................... | .................... | .................... |
| X | 2807 hCoV-19/Belgium/ULG-11044/2020\|EPI_ISL_722957\|2020 | .................... | .................... | .................... | .................... | .................... | .................... |
| X | 2808 hCoV-19/Belgium/ULG-11045/2020\|EPI_ISL_722958\|2020 | .................... | .................... | .................... | .................... | .................... | .................... |
| X | 2809 hCoV-19/Belgium/ULG-11048/2020\|EPI_ISL_722959\|2020 | .................... | .................... | .................... | .................... | ......T............. | .................... |
| X | 2810 hCoV-19/Belgium/ULG-11050/2020\|EPI_ISL_722960\|2020 | .................... | .................... | .................... | .................... | ......T............. | .................... |
| X | 2811 hCoV-19/Belgium/ULG-11051/2020\|EPI_ISL_722961\|2020 | .................... | .................... | .................... | .................... | .................... | .................... |
| X | 2812 hCoV-19/Belgium/ULG-11052/2020\|EPI_ISL_722962\|2020 | .................... | .................... | .................... | .................... | .................... | .................... |
| X | 2813 hCoV-19/Belgium/ULG-11053/2020\|EPI_ISL_722963\|2020 | .................... | .................... | .................... | .................... | .................... | .................... |
| X | 2814 hCoV-19/Belgium/ULG-11054/2020\|EPI_ISL_722964\|2020 | .................... | .................... | .................... | .................... | .................... | .................... |
| X | 2815 hCoV-19/Belgium/ULG-11055/2020\|EPI_ISL_722965\|2020 | .................... | .................... | .................... | .................... | .................... | .................... |

| | | | | | | | | |
|---|---|---|---|---|---|---|---|---|
| X | 2816 | hCoV-19/Belgium/ULG-11057/2020\|EPI_ISL_722966\|2020 | ···················· | ·A·················· | ···················· | ···················· | ···················· | ···················· |
| X | 2817 | hCoV-19/Denmark/DCGC-15263/2020\|EPI_ISL_713270\|202 | ···················· | ···················· | ···················· | ···················· | ···················· | ···················· |
| X | 2818 | hCoV-19/Denmark/DCGC-15898/2020\|EPI_ISL_713271\|202 | ···················· | ···················· | ···················· | ···················· | ···················· | ···················· |
| X | 2819 | hCoV-19/Denmark/DCGC-15900/2020\|EPI_ISL_713272\|202 | ···················· | ···················· | ···················· | ···················· | ···················· | ···················· |
| X | 2820 | hCoV-19/Denmark/DCGC-14942/2020\|EPI_ISL_713273\|202 | ···················· | ···················· | ···················· | ···················· | ···················· | ···················· |
| X | 2821 | hCoV-19/Denmark/DCGC-14983/2020\|EPI_ISL_713274\|202 | ···················· | ···················· | ···················· | ···················· | ···················· | ···················· |
| X | 2822 | hCoV-19/Denmark/DCGC-14992/2020\|EPI_ISL_713275\|202 | ···················· | ···················· | ···················· | ···················· | ···················· | ···················· |
| X | 2823 | hCoV-19/Denmark/DCGC-15012/2020\|EPI_ISL_713276\|202 | ···················· | ···················· | ···················· | ···················· | ···················· | ···················· |
| X | 2824 | hCoV-19/Denmark/DCGC-15078/2020\|EPI_ISL_713277\|202 | ···················· | ···················· | ···················· | ···················· | ···················· | ···················· |
| X | 2825 | hCoV-19/Denmark/DCGC-15079/2020\|EPI_ISL_713278\|202 | ···················· | ···················· | ···················· | ···················· | ···················· | ···················· |
| X | 2826 | hCoV-19/Denmark/DCGC-15082/2020\|EPI_ISL_713279\|202 | ···················· | ···················· | ···················· | ···················· | ···················· | ···················· |
| X | 2827 | hCoV-19/Denmark/DCGC-15126/2020\|EPI_ISL_713280\|202 | ···················· | ···················· | ···················· | ···················· | ···················· | ···················· |
| X | 2828 | hCoV-19/Denmark/DCGC-15138/2020\|EPI_ISL_713281\|202 | ···················· | ···················· | ···················· | ···················· | ···················· | ···················· |
| X | 2829 | hCoV-19/Denmark/DCGC-15163/2020\|EPI_ISL_713282\|202 | ···················· | ···················· | ···················· | ···················· | ···················· | ···················· |
| X | 2830 | hCoV-19/Denmark/DCGC-15167/2020\|EPI_ISL_713283\|202 | ···················· | ···················· | ···················· | ···················· | ···················· | ···················· |
| X | 2831 | hCoV-19/Denmark/DCGC-15236/2020\|EPI_ISL_713284\|202 | ···················· | ···················· | ···················· | ···················· | ···················· | ···················· |
| X | 2832 | hCoV-19/Denmark/DCGC-15292/2020\|EPI_ISL_713285\|202 | ···················· | ···················· | ···················· | ···················· | ···················· | ···················· |
| X | 2833 | hCoV-19/Denmark/DCGC-15329/2020\|EPI_ISL_713286\|202 | ···················· | ···················· | ···················· | ···················· | ···················· | ···················· |
| X | 2834 | hCoV-19/Denmark/DCGC-15490/2020\|EPI_ISL_713287\|202 | ···················· | ···················· | ···················· | ···················· | ···················· | ···················· |
| X | 2835 | hCoV-19/Denmark/DCGC-15539/2020\|EPI_ISL_713288\|202 | ···················· | ···················· | ···················· | ···················· | ···················· | ···················· |
| X | 2836 | hCoV-19/Denmark/DCGC-15579/2020\|EPI_ISL_713289\|202 | ···················· | ···················· | ···················· | ···················· | ···················· | ···················· |
| X | 2837 | hCoV-19/Denmark/DCGC-15499/2020\|EPI_ISL_713291\|202 | ···················· | ···················· | ···················· | ···················· | ···················· | ···················· |
| X | 2838 | hCoV-19/Denmark/DCGC-15623/2020\|EPI_ISL_713292\|202 | ···················· | ···················· | ···················· | ···················· | ···················· | ···················· |
| X | 2839 | hCoV-19/Denmark/DCGC-15077/2020\|EPI_ISL_713293\|202 | ···················· | ···················· | ···················· | ···················· | ···················· | ···················· |
| X | 2840 | hCoV-19/Denmark/DCGC-15265/2020\|EPI_ISL_713294\|202 | ···················· | ···················· | ···················· | ···················· | ···················· | ···················· |
| X | 2841 | hCoV-19/Denmark/DCGC-15949/2020\|EPI_ISL_714500\|202 | ···················· | ···················· | ···················· | ···················· | ···················· | ···················· |
| X | 2842 | hCoV-19/Denmark/DCGC-15950/2020\|EPI_ISL_714501\|202 | ···················· | ···················· | ···················· | ···················· | ···················· | ···················· |
| X | 2843 | hCoV-19/Denmark/DCGC-15951/2020\|EPI_ISL_714502\|202 | ···················· | ···················· | ···················· | ···················· | ···················· | ···················· |
| X | 2844 | hCoV-19/Denmark/DCGC-15953/2020\|EPI_ISL_714503\|202 | ···················· | ···················· | ···················· | ···················· | ···················· | ···················· |
| X | 2845 | hCoV-19/Denmark/DCGC-15954/2020\|EPI_ISL_714504\|202 | ···················· | ···················· | ···················· | ···················· | ···················· | ···················· |
| X | 2846 | hCoV-19/Denmark/DCGC-15956/2020\|EPI_ISL_714505\|202 | ···················· | ···················· | ···················· | ···················· | ···················· | ···················· |
| X | 2847 | hCoV-19/Denmark/DCGC-15959/2020\|EPI_ISL_714506\|202 | ···················· | ···················· | ···················· | ···················· | ···················· | ···················· |
| X | 2848 | hCoV-19/Denmark/DCGC-15962/2020\|EPI_ISL_714507\|202 | ···················· | ···················· | ···················· | ···················· | ···················· | ···················· |
| X | 2849 | hCoV-19/Denmark/DCGC-15963/2020\|EPI_ISL_714508\|202 | ···················· | ···················· | ···················· | ···················· | ·····T·············· | ···················· |
| X | 2850 | hCoV-19/Denmark/DCGC-15965/2020\|EPI_ISL_714509\|202 | ···················· | ···················· | ···················· | ···················· | ···················· | ···················· |
| X | 2851 | hCoV-19/Denmark/DCGC-15967/2020\|EPI_ISL_714510\|202 | ···················· | ···················· | ···················· | ···················· | ···················· | ···················· |
| X | 2852 | hCoV-19/Denmark/DCGC-15969/2020\|EPI_ISL_714511\|202 | ···················· | ···················· | ···················· | ···················· | ···················· | ···················· |
| X | 2853 | hCoV-19/Denmark/DCGC-15971/2020\|EPI_ISL_714512\|202 | ···················· | ·A·················· | ···················· | ···················· | ···················· | ··A················· |
| X | 2854 | hCoV-19/Denmark/DCGC-15973/2020\|EPI_ISL_714513\|202 | ···················· | ···················· | ···················· | ···················· | ···················· | ···················· |
| X | 2855 | hCoV-19/Denmark/DCGC-15974/2020\|EPI_ISL_714514\|202 | ···················· | ···················· | ···················· | ···················· | ···················· | ···················· |
| X | 2856 | hCoV-19/Denmark/DCGC-15975/2020\|EPI_ISL_714515\|202 | ···················· | ···················· | ···················· | ···················· | ···················· | ···················· |
| X | 2857 | hCoV-19/Denmark/DCGC-15980/2020\|EPI_ISL_714516\|202 | ···················· | ···················· | ···················· | ···················· | ···················· | ···················· |
| X | 2858 | hCoV-19/Denmark/DCGC-15985/2020\|EPI_ISL_714517\|202 | ···················· | ···················· | ···················· | ···················· | ···················· | ···················· |
| X | 2859 | hCoV-19/Denmark/DCGC-15987/2020\|EPI_ISL_714518\|202 | ···················· | ···················· | ···················· | ···················· | ···················· | ···················· |
| X | 2860 | hCoV-19/Denmark/DCGC-15989/2020\|EPI_ISL_714519\|202 | ···················· | ···················· | ···················· | ···················· | ···················· | ···················· |
| X | 2861 | hCoV-19/Denmark/DCGC-15990/2020\|EPI_ISL_714520\|202 | ···················· | ···················· | ···················· | ···················· | ···················· | ···················· |
| X | 2862 | hCoV-19/Denmark/DCGC-15993/2020\|EPI_ISL_714521\|202 | ···················· | ···················· | ···················· | ···················· | ···················· | ···················· |
| X | 2863 | hCoV-19/Denmark/DCGC-15994/2020\|EPI_ISL_714522\|202 | ···················· | ···················· | ···················· | ···················· | ·····T·············· | ···················· |
| X | 2864 | hCoV-19/Denmark/DCGC-15997/2020\|EPI_ISL_714523\|202 | ···················· | ···················· | ···················· | ···················· | ·····T·············· | ···················· |
| X | 2865 | hCoV-19/Denmark/DCGC-15998/2020\|EPI_ISL_714524\|202 | ···················· | ·A·················· | ···················· | ···················· | ···················· | ··A················· |
| X | 2866 | hCoV-19/Denmark/DCGC-16000/2020\|EPI_ISL_714525\|202 | ···················· | ···················· | ···················· | ···················· | ···················· | ···················· |
| X | 2867 | hCoV-19/Denmark/DCGC-16001/2020\|EPI_ISL_714526\|202 | ···················· | ·A·················· | ···················· | ···················· | ···················· | ··A················· |
| X | 2868 | hCoV-19/Denmark/DCGC-16002/2020\|EPI_ISL_714527\|202 | ···················· | ···················· | ···················· | ···················· | ···················· | ···················· |
| X | 2869 | hCoV-19/Denmark/DCGC-16003/2020\|EPI_ISL_714528\|202 | ···················· | ···················· | ···················· | ···················· | ···················· | ···················· |
| X | 2870 | hCoV-19/Denmark/DCGC-16004/2020\|EPI_ISL_714529\|202 | ···················· | ···················· | ···················· | ···················· | ···················· | ···················· |
| X | 2871 | hCoV-19/Denmark/DCGC-16006/2020\|EPI_ISL_714530\|202 | ···················· | ···················· | ···················· | ···················· | ···················· | ···················· |
| X | 2872 | hCoV-19/Denmark/DCGC-16007/2020\|EPI_ISL_714531\|202 | ···················· | ···················· | ···················· | ···················· | ···················· | ···················· |
| X | 2873 | hCoV-19/Denmark/DCGC-16010/2020\|EPI_ISL_714532\|202 | ···················· | ·A·················· | ···················· | ···················· | ···················· | ··A················· |
| X | 2874 | hCoV-19/Denmark/DCGC-16011/2020\|EPI_ISL_714533\|202 | ···················· | ···················· | ···················· | ···················· | ···················· | ···················· |
| X | 2875 | hCoV-19/Denmark/DCGC-16012/2020\|EPI_ISL_714534\|202 | ···················· | ···················· | ···················· | ···················· | ···················· | ···················· |
| X | 2876 | hCoV-19/Denmark/DCGC-16013/2020\|EPI_ISL_714535\|202 | ···················· | ···················· | ···················· | ···················· | ···················· | ···················· |
| X | 2877 | hCoV-19/Denmark/DCGC-16016/2020\|EPI_ISL_714536\|202 | ···················· | ···················· | ···················· | ···················· | ···················· | ···················· |
| X | 2878 | hCoV-19/Denmark/DCGC-16017/2020\|EPI_ISL_714537\|202 | ···················· | ···················· | ···················· | ···················· | ···················· | ···················· |
| X | 2879 | hCoV-19/Denmark/DCGC-16018/2020\|EPI_ISL_714538\|202 | ···················· | ···················· | ···················· | ···················· | ···················· | ···················· |
| X | 2880 | hCoV-19/Denmark/DCGC-16019/2020\|EPI_ISL_714539\|202 | ···················· | ···················· | ···················· | ···················· | ···················· | ···················· |
| X | 2881 | hCoV-19/Denmark/DCGC-16020/2020\|EPI_ISL_714540\|202 | ···················· | ···················· | ···················· | ···················· | ···················· | ···················· |
| X | 2882 | hCoV-19/Denmark/DCGC-16021/2020\|EPI_ISL_714541\|202 | ···················· | ·A·················· | ···················· | ···················· | ···················· | ···················· |
| X | 2883 | hCoV-19/Denmark/DCGC-16022/2020\|EPI_ISL_714542\|202 | ···················· | ···················· | ···················· | ···················· | ···················· | ···················· |
| X | 2884 | hCoV-19/Denmark/DCGC-16024/2020\|EPI_ISL_714543\|202 | ···················· | ···················· | ···················· | ···················· | ···················· | ···················· |
| X | 2885 | hCoV-19/Denmark/DCGC-16025/2020\|EPI_ISL_714544\|202 | ···················· | ···················· | ···················· | ···················· | ·····T·············· | ···················· |
| X | 2886 | hCoV-19/Denmark/DCGC-16026/2020\|EPI_ISL_714545\|202 | ···················· | ···················· | ···················· | ···················· | ···················· | ···················· |
| X | 2887 | hCoV-19/Denmark/DCGC-16030/2020\|EPI_ISL_714546\|202 | ···················· | ···················· | ···················· | ···················· | ···················· | ···················· |
| X | 2888 | hCoV-19/Denmark/DCGC-16031/2020\|EPI_ISL_714547\|202 | ···················· | ···················· | ···················· | ···················· | ···················· | ···················· |
| X | 2889 | hCoV-19/Denmark/DCGC-16032/2020\|EPI_ISL_714548\|202 | ···················· | ·A·················· | ···················· | ···················· | ···················· | ···················· |
| X | 2890 | hCoV-19/Denmark/DCGC-16033/2020\|EPI_ISL_714549\|202 | ···················· | ···················· | ···················· | ···················· | ···················· | ···················· |
| X | 2891 | hCoV-19/Denmark/DCGC-16034/2020\|EPI_ISL_714550\|202 | ···················· | ·A·················· | ···················· | ···················· | ···················· | ···················· |
| X | 2892 | hCoV-19/Denmark/DCGC-16040/2020\|EPI_ISL_714551\|202 | ···················· | ···················· | ···················· | ···················· | ···················· | ···················· |
| X | 2893 | hCoV-19/Denmark/DCGC-16041/2020\|EPI_ISL_714552\|202 | ···················· | ···················· | ···················· | ···················· | ···················· | ···················· |
| X | 2894 | hCoV-19/Denmark/DCGC-16042/2020\|EPI_ISL_714553\|202 | ···················· | ···················· | ···················· | ···················· | ···················· | ···················· |
| X | 2895 | hCoV-19/Denmark/DCGC-16044/2020\|EPI_ISL_714554\|202 | ···················· | ···················· | ···················· | ···················· | ···················· | ···················· |

| | | | | | | | |
|---|---|---|---|---|---|---|---|
| X | 2896 hCoV-19/Denmark/DCGC-16045/2020\|EPI_ISL_714555\|202 | ···················· | ···················· | ···················· | ···················· | ···················· | ···················· |
| X | 2897 hCoV-19/Denmark/DCGC-16046/2020\|EPI_ISL_714556\|202 | ···················· | ···················· | ···················· | ···················· | ···················· | ···················· |
| X | 2898 hCoV-19/Denmark/DCGC-16048/2020\|EPI_ISL_714557\|202 | ···················· | ···················· | ···················· | ···················· | ···················· | ···················· |
| X | 2899 hCoV-19/Denmark/DCGC-16050/2020\|EPI_ISL_714558\|202 | ···················· | ···················· | ···················· | ···················· | ······T············· | ···················· |
| X | 2900 hCoV-19/Denmark/DCGC-16051/2020\|EPI_ISL_714559\|202 | ···················· | ···················· | ···················· | ···················· | ···················· | ···················· |
| X | 2901 hCoV-19/Denmark/DCGC-16052/2020\|EPI_ISL_714560\|202 | ···················· | ···················· | ···················· | ···················· | ···················· | ···················· |
| X | 2902 hCoV-19/Denmark/DCGC-16053/2020\|EPI_ISL_714561\|202 | ···················· | ···················· | ···················· | ···················· | ···················· | ···················· |
| X | 2903 hCoV-19/Denmark/DCGC-16054/2020\|EPI_ISL_714562\|202 | ···················· | ···················· | ···················· | ···················· | ···················· | ···················· |
| X | 2904 hCoV-19/Denmark/DCGC-16055/2020\|EPI_ISL_714563\|202 | ···················· | ···················· | ···················· | ···················· | ···················· | ···················· |
| X | 2905 hCoV-19/Denmark/DCGC-16056/2020\|EPI_ISL_714564\|202 | ···················· | ···················· | ···················· | ···················· | ···················· | ···················· |
| X | 2906 hCoV-19/Denmark/DCGC-16057/2020\|EPI_ISL_714565\|202 | ···················· | ···················· | ···················· | ···················· | ······T············· | ···················· |
| X | 2907 hCoV-19/Denmark/DCGC-16059/2020\|EPI_ISL_714566\|202 | ···················· | ···················· | ···················· | ···················· | ···················· | ···················· |
| X | 2908 hCoV-19/Denmark/DCGC-16060/2020\|EPI_ISL_714567\|202 | ···················· | ·A·················· | ···················· | ···················· | ···················· | ···A················ |
| X | 2909 hCoV-19/Denmark/DCGC-16061/2020\|EPI_ISL_714568\|202 | ···················· | ·A·················· | ·········T·········· | ···················· | ···················· | ···················· |
| X | 2910 hCoV-19/Denmark/DCGC-16065/2020\|EPI_ISL_714569\|202 | ···················· | ···················· | ···················· | ···················· | ······T············· | ···················· |
| X | 2911 hCoV-19/Denmark/DCGC-16067/2020\|EPI_ISL_714570\|202 | ···················· | ···················· | ···················· | ···················· | ···················· | ···················· |
| X | 2912 hCoV-19/Denmark/DCGC-16069/2020\|EPI_ISL_714571\|202 | ···················· | ···················· | ···················· | ···················· | ···················· | ···················· |
| X | 2913 hCoV-19/Denmark/DCGC-16070/2020\|EPI_ISL_714572\|202 | ···················· | ···················· | ···················· | ···················· | ···················· | ···················· |
| X | 2914 hCoV-19/Denmark/DCGC-16071/2020\|EPI_ISL_714573\|202 | ···················· | ···················· | ···················· | ···················· | ···················· | ···················· |
| X | 2915 hCoV-19/Denmark/DCGC-16072/2020\|EPI_ISL_714574\|202 | ···················· | ·A·················· | ···················· | ···················· | ···················· | ···················· |
| X | 2916 hCoV-19/Denmark/DCGC-16076/2020\|EPI_ISL_714575\|202 | ···················· | ···················· | ···················· | ···················· | ···················· | ···················· |
| X | 2917 hCoV-19/Denmark/DCGC-16083/2020\|EPI_ISL_714576\|202 | ···················· | ···················· | ···················· | ···················· | ···················· | ···················· |
| X | 2918 hCoV-19/Denmark/DCGC-16084/2020\|EPI_ISL_714577\|202 | ···················· | ···················· | ···················· | ···················· | ···················· | ···················· |
| X | 2919 hCoV-19/Denmark/DCGC-16085/2020\|EPI_ISL_714578\|202 | ···················· | ···················· | ···················· | ···················· | ···················· | ···················· |
| X | 2920 hCoV-19/Denmark/DCGC-16087/2020\|EPI_ISL_714579\|202 | ···················· | ···················· | ···················· | ···················· | ···················· | ···················· |
| X | 2921 hCoV-19/Denmark/DCGC-16090/2020\|EPI_ISL_714580\|202 | ···················· | ···················· | ···················· | ···················· | ···················· | ···················· |
| X | 2922 hCoV-19/Denmark/DCGC-16092/2020\|EPI_ISL_714581\|202 | ···················· | ···················· | ···················· | ···················· | ···················· | ···················· |
| X | 2923 hCoV-19/Denmark/DCGC-16094/2020\|EPI_ISL_714582\|202 | ···················· | ···················· | ···················· | ···················· | ···················· | ···················· |
| X | 2924 hCoV-19/Denmark/DCGC-16095/2020\|EPI_ISL_714583\|202 | ···················· | ···················· | ···················· | ···················· | ···················· | ···················· |
| X | 2925 hCoV-19/Denmark/DCGC-16096/2020\|EPI_ISL_714584\|202 | ···················· | ···················· | ···················· | ···················· | ···················· | ···················· |
| X | 2926 hCoV-19/Denmark/DCGC-16098/2020\|EPI_ISL_714585\|202 | ···················· | ···················· | ···················· | ···················· | ···················· | ···················· |
| X | 2927 hCoV-19/Denmark/DCGC-16106/2020\|EPI_ISL_714586\|202 | ···················· | ···················· | ···················· | ···················· | ···················· | ···················· |
| X | 2928 hCoV-19/Denmark/DCGC-16107/2020\|EPI_ISL_714587\|202 | ···················· | ···················· | ···················· | ···················· | ···················· | ···················· |
| X | 2929 hCoV-19/Denmark/DCGC-16109/2020\|EPI_ISL_714588\|202 | ···················· | ···················· | ···················· | ···················· | ···················· | ···················· |
| X | 2930 hCoV-19/Denmark/DCGC-16113/2020\|EPI_ISL_714589\|202 | ···················· | ···················· | ···················· | ···················· | ···················· | ···················· |
| X | 2931 hCoV-19/Denmark/DCGC-16115/2020\|EPI_ISL_714590\|202 | ···················· | ···················· | ···················· | ···················· | ···················· | ···················· |
| X | 2932 hCoV-19/Denmark/DCGC-16116/2020\|EPI_ISL_714591\|202 | ···················· | ···················· | ···················· | ···················· | ···················· | ···················· |
| X | 2933 hCoV-19/Denmark/DCGC-16118/2020\|EPI_ISL_714592\|202 | ···················· | ···················· | ···················· | ···················· | ···················· | ···················· |
| X | 2934 hCoV-19/Denmark/DCGC-16119/2020\|EPI_ISL_714593\|202 | ···················· | ···················· | ···················· | ···················· | ···················· | ···················· |
| X | 2935 hCoV-19/Denmark/DCGC-16120/2020\|EPI_ISL_714594\|202 | ···················· | ···················· | ···················· | ···················· | ···················· | ···················· |
| X | 2936 hCoV-19/Denmark/DCGC-16121/2020\|EPI_ISL_714595\|202 | ···················· | ···················· | ···················· | ···················· | ···················· | ···················· |
| X | 2937 hCoV-19/Denmark/DCGC-16122/2020\|EPI_ISL_714596\|202 | ···················· | ···················· | ···················· | ···················· | ···················· | ···················· |
| X | 2938 hCoV-19/Denmark/DCGC-16124/2020\|EPI_ISL_714597\|202 | ···················· | ···················· | ···················· | ···················· | ······T············· | ···················· |
| X | 2939 hCoV-19/Denmark/DCGC-16125/2020\|EPI_ISL_714598\|202 | ···················· | ····A··············· | ···················· | ···················· | ···················· | ···················· |
| X | 2940 hCoV-19/Denmark/DCGC-16128/2020\|EPI_ISL_714599\|202 | ···················· | ···················· | ···················· | ···················· | ······T············· | ···················· |
| X | 2941 hCoV-19/Germany/NW-HHU-329/2020\|EPI_ISL_718031\|202 | ···················· | ···················· | ···················· | ···················· | ···················· | ···················· |
| X | 2942 hCoV-19/Germany/NW-HHU-330/2020\|EPI_ISL_718032\|202 | ···················· | ···················· | ···················· | ···················· | ···················· | ···················· |
| X | 2943 hCoV-19/Germany/NW-HHU-389/2020\|EPI_ISL_718070\|202 | ···················· | ···················· | ···················· | ···················· | ···················· | ···················· |
| X | 2944 hCoV-19/Germany/NW-HHU-390/2020\|EPI_ISL_718071\|202 | ···················· | ···················· | ···················· | ···················· | ···················· | ···················· |
| X | 2945 hCoV-19/Germany/NW-HHU-391/2020\|EPI_ISL_718072\|202 | ···················· | ···················· | ···················· | ···················· | ···················· | ···················· |
| X | 2946 hCoV-19/Germany/NW-HHU-392/2020\|EPI_ISL_718073\|202 | ···················· | ···················· | ···················· | ···················· | ···················· | ···················· |
| X | 2947 hCoV-19/Germany/NW-HHU-394/2020\|EPI_ISL_718074\|202 | ···················· | ···················· | ···················· | ···················· | ···················· | ···················· |
| X | 2948 hCoV-19/Germany/NW-HHU-395/2020\|EPI_ISL_718075\|202 | ···················· | ···················· | ···················· | ···················· | ···················· | ···················· |
| X | 2949 hCoV-19/Germany/NW-HHU-396/2020\|EPI_ISL_718076\|202 | ···················· | ···················· | ···················· | ···················· | ···················· | ···················· |
| X | 2950 hCoV-19/Germany/NW-HHU-398/2020\|EPI_ISL_718077\|202 | ···················· | ···················· | ···················· | ···················· | ···················· | ···················· |
| X | 2951 hCoV-19/Germany/NW-HHU-399/2020\|EPI_ISL_718078\|202 | ···················· | ···················· | ···················· | ···················· | ···················· | ···················· |
| X | 2952 hCoV-19/Germany/NW-HHU-401/2020\|EPI_ISL_718079\|202 | ···················· | ·········A·········· | ···················· | ···················· | ···················· | ···················· |
| X | 2953 hCoV-19/Germany/NW-HHU-404/2020\|EPI_ISL_718080\|202 | ···················· | ···················· | ···················· | ···················· | ···················· | ···················· |
| X | 2954 hCoV-19/Germany/NW-HHU-405/2020\|EPI_ISL_718081\|202 | ···················· | ···················· | ···················· | ···················· | ···················· | ···················· |
| X | 2955 hCoV-19/Germany/NW-HHU-406/2020\|EPI_ISL_718082\|202 | ···················· | ···················· | ···················· | ···················· | ···················· | ················A··· |
| X | 2956 hCoV-19/Germany/NW-HHU-409/2020\|EPI_ISL_718083\|202 | ···················· | ···················· | ···················· | ···················· | ···················· | ···················· |
| X | 2957 hCoV-19/Germany/NW-HHU-410/2020\|EPI_ISL_718084\|202 | ···················· | ···················· | ···················· | ···················· | ···················· | ···················· |
| X | 2958 hCoV-19/Germany/NW-HHU-413/2020\|EPI_ISL_718087\|202 | ···················· | ······A············· | ···················· | ···················· | ···················· | ···················· |
| X | 2959 hCoV-19/Germany/NW-HHU-414/2020\|EPI_ISL_718088\|202 | ···················· | ···················· | ···················· | ···················· | ···················· | ···················· |
| X | 2960 hCoV-19/Germany/NW-HHU-415/2020\|EPI_ISL_718089\|202 | ···················· | ······A············· | ···················· | ···················· | ···················· | ···················· |
| X | 2961 hCoV-19/Germany/NW-HHU-416/2020\|EPI_ISL_718090\|202 | ···················· | ···················· | ···················· | ···················· | ···················· | ···················· |
| X | 2962 hCoV-19/Germany/NW-HHU-417/2020\|EPI_ISL_718091\|202 | ···················· | ···················· | ···················· | ···················· | ···················· | ···················· |
| X | 2963 hCoV-19/Germany/NW-HHU-418/2020\|EPI_ISL_718092\|202 | ···················· | ···················· | ···················· | ···················· | ···················· | ···················· |
| X | 2964 hCoV-19/Germany/NW-HHU-419/2020\|EPI_ISL_718093\|202 | ···················· | ···················· | ···················· | ···················· | ···················· | ···················· |
| X | 2965 hCoV-19/Germany/NW-HHU-421/2020\|EPI_ISL_718095\|202 | ···················· | ···················· | ···················· | ···················· | ···················· | ···················· |
| X | 2966 hCoV-19/Germany/NW-HHU-423/2020\|EPI_ISL_718097\|202 | ···················· | ···················· | ···················· | ···················· | ···················· | ···················· |
| X | 2967 hCoV-19/Germany/NW-HHU-424/2020\|EPI_ISL_718098\|202 | ···················· | ···················· | ···················· | ···················· | ···················· | ···················· |
| X | 2968 hCoV-19/Germany/NW-HHU-425/2020\|EPI_ISL_718099\|202 | ···················· | ·A·················· | ···················· | ···················· | ···················· | ···················· |
| X | 2969 hCoV-19/Germany/NW-HHU-426/2020\|EPI_ISL_718100\|202 | ···················· | ···················· | ···················· | ···················· | ···················· | ···················· |
| X | 2970 hCoV-19/Germany/NW-HHU-427/2020\|EPI_ISL_718101\|202 | ···················· | ·A·················· | ···················· | ···················· | ···················· | ···················· |
| X | 2971 hCoV-19/Germany/NW-HHU-430/2020\|EPI_ISL_718104\|202 | ···················· | ···················· | ···················· | ···················· | ···················· | ···················· |
| X | 2972 hCoV-19/England/CAMC-C43F71/2020\|EPI_ISL_719370\|20 | ···················· | ·A·················· | ···················· | ···················· | ···················· | ···················· |
| X | 2973 hCoV-19/England/CAMC-C44866/2020\|EPI_ISL_719372\|20 | ···················· | ·A·················· | ···················· | ···················· | ···················· | ···A················ |
| X | 2974 hCoV-19/England/CAMC-C432D2/2020\|EPI_ISL_719375\|20 | ···················· | ···················· | ···················· | ···················· | ···················· | ···················· |
| X | 2975 hCoV-19/England/CAMC-C45883/2020\|EPI_ISL_719377\|20 | ···················· | ···················· | ···················· | ···················· | ···················· | ···················· |

| | | | | | | | | |
|---|---|---|---|---|---|---|---|---|
| X | 2976 | hCoV-19/England/CAMC-C4423B/2020\|EPI_ISL_719407\|20 | ···················· | ···················· | ····················· | ···················· | ···················· | ···················· |
| X | 2977 | hCoV-19/England/CAMC-C447C3/2020\|EPI_ISL_719416\|20 | ···················· | ···················· | ····················· | ···················· | ···················· | ···················· |
| X | 2978 | hCoV-19/England/CAMC-C43946/2020\|EPI_ISL_719422\|20 | ···················· | ···················· | ····················· | ···················· | ···················· | ···················· |
| X | 2979 | hCoV-19/England/CAMC-C456B6/2020\|EPI_ISL_719425\|20 | ···················· | ···················· | ····················· | ···················· | ···················· | ···················· |
| X | 2980 | hCoV-19/England/CAMC-C4409B/2020\|EPI_ISL_719428\|20 | ···················· | ···················· | ····················· | ···················· | ···················· | ···················· |
| X | 2981 | hCoV-19/England/MILK-BB0F07/2020\|EPI_ISL_720359\|20 | ···················· | ···················· | ····················· | ···················· | ···················· | ···················· |
| X | 2982 | hCoV-19/England/MILK-BAC548/2020\|EPI_ISL_720360\|20 | ···················· | ···················· | ····················· | ···················· | ···················· | ···················· |
| X | 2983 | hCoV-19/England/MILK-BB116A/2020\|EPI_ISL_720361\|20 | ···················· | ···················· | ····················· | ···················· | ······T············· | ···················· |
| X | 2984 | hCoV-19/England/MILK-BB1179/2020\|EPI_ISL_720362\|20 | ···················· | ·A·················· | ····················· | ···················· | ···················· | ···A················ |
| X | 2985 | hCoV-19/England/MILK-BB108B/2020\|EPI_ISL_720363\|20 | ···················· | ···················· | ····················· | ···················· | ···················· | ···················· |
| X | 2986 | hCoV-19/England/MILK-BB005F/2020\|EPI_ISL_720365\|20 | ···················· | ···················· | ····················· | ···················· | ···················· | ···················· |
| X | 2987 | hCoV-19/England/MILK-BB0EBF/2020\|EPI_ISL_720366\|20 | ···················· | ···················· | ····················· | ···················· | ···················· | ···················· |
| X | 2988 | hCoV-19/England/MILK-BAFBF8/2020\|EPI_ISL_720368\|20 | ···················· | ···················· | ····················· | ···················· | ···················· | ···················· |
| X | 2989 | hCoV-19/England/MILK-BB0C00/2020\|EPI_ISL_720390\|20 | ···················· | ···················· | ····················· | ···················· | ···················· | ···················· |
| X | 2990 | hCoV-19/England/MILK-BABA86/2020\|EPI_ISL_720392\|20 | ···················· | ···················· | ····················· | ···················· | ···················· | ···················· |
| X | 2991 | hCoV-19/England/MILK-BAC821/2020\|EPI_ISL_720393\|20 | ···················· | ···················· | ····················· | ···················· | ···················· | ···················· |
| X | 2992 | hCoV-19/England/MILK-BAC320/2020\|EPI_ISL_720395\|20 | ···················· | ···················· | ····················· | ···················· | ······T············· | ···················· |
| X | 2993 | hCoV-19/England/MILK-BB0EDD/2020\|EPI_ISL_720396\|20 | ···················· | ···················· | ····················· | ···················· | ···················· | ···················· |
| X | 2994 | hCoV-19/England/MILK-BABA3B/2020\|EPI_ISL_720397\|20 | ···················· | ···················· | ····················· | ···················· | ···················· | ···················· |
| X | 2995 | hCoV-19/England/MILK-BB0022/2020\|EPI_ISL_720398\|20 | ···················· | ···················· | ····················· | ···················· | ···················· | ···················· |
| X | 2996 | hCoV-19/England/MILK-BB2336/2020\|EPI_ISL_720399\|20 | ···················· | ···················· | ····················· | ···················· | ···················· | ···················· |
| X | 2997 | hCoV-19/England/MILK-BAC52A/2020\|EPI_ISL_720400\|20 | ···················· | ···················· | ····················· | ···················· | ···················· | ···················· |
| X | 2998 | hCoV-19/England/MILK-BB0A06/2020\|EPI_ISL_720401\|20 | ···················· | ·A·················· | ····················· | ···················· | ···················· | ···A················ |
| X | 2999 | hCoV-19/England/MILK-BB2318/2020\|EPI_ISL_720404\|20 | ···················· | ···················· | ····················· | ···················· | ···················· | ···················· |
| X | 3000 | hCoV-19/England/MILK-BABB56/2020\|EPI_ISL_720405\|20 | ···················· | ···················· | ····················· | ···················· | ···················· | ···················· |
| X | 3001 | hCoV-19/England/MILK-BB0EFB/2020\|EPI_ISL_720406\|20 | ···················· | ···················· | ····················· | ···················· | ···················· | ···················· |
| X | 3002 | hCoV-19/England/MILK-BB2372/2020\|EPI_ISL_720407\|20 | ···················· | ···················· | ····················· | ···················· | ···················· | ···················· |
| X | 3003 | hCoV-19/England/MILK-BB0356/2020\|EPI_ISL_720409\|20 | ···················· | ·A·················· | ····················· | ···················· | ···················· | ···················· |
| X | 3004 | hCoV-19/England/MILK-BABA59/2020\|EPI_ISL_720410\|20 | ···················· | ···················· | ····················· | ···················· | ···················· | ···················· |
| X | 3005 | hCoV-19/England/MILK-BB22A2/2020\|EPI_ISL_720411\|20 | ···················· | ···················· | ····················· | ···················· | ···················· | ···················· |
| X | 3006 | hCoV-19/England/MILK-BB0C79/2020\|EPI_ISL_720412\|20 | ···················· | ···················· | ····················· | ···················· | ···················· | ···················· |
| X | 3007 | hCoV-19/England/MILK-BB065D/2020\|EPI_ISL_720413\|20 | ···················· | ···················· | ····················· | ···················· | ···················· | ···················· |
| X | 3008 | hCoV-19/England/MILK-BB0383/2020\|EPI_ISL_720418\|20 | ···················· | ···················· | ····················· | ···················· | ···················· | ···················· |
| X | 3009 | hCoV-19/England/MILK-BAFC04/2020\|EPI_ISL_720421\|20 | ···················· | ···················· | ····················· | ···················· | ···················· | ···················· |
| X | 3010 | hCoV-19/England/MILK-BB0E28/2020\|EPI_ISL_720423\|20 | ···················· | ···················· | ····················· | ···················· | ···················· | ···················· |
| X | 3011 | hCoV-19/England/MILK-BB0E73/2020\|EPI_ISL_720424\|20 | ···················· | ···················· | ····················· | ···················· | ···················· | ···················· |
| X | 3012 | hCoV-19/Switzerland/SO-ETHZ-390228/2020\|EPI_ISL_72 | ···················· | ···················· | ····················· | ···················· | ······K············· | ···················· |
| X | 3013 | hCoV-19/Switzerland/BL-ETHZ-380270/2020\|EPI_ISL_72 | ···················· | ···················· | ····················· | ···················· | ···················· | ···················· |
| X | 3014 | hCoV-19/Switzerland/AG-ETHZ-380325/2020\|EPI_ISL_72 | ···················· | ···················· | ····················· | ···················· | ···················· | ···················· |
| X | 3015 | hCoV-19/Switzerland/AG-ETHZ-380374/2020\|EPI_ISL_72 | ···················· | ···················· | ····················· | ···················· | ···················· | ···················· |
| X | 3016 | hCoV-19/Switzerland/SG-ETHZ-390223/2020\|EPI_ISL_72 | ···················· | ···················· | ····················· | ···················· | ···················· | ···················· |
| X | 3017 | hCoV-19/Switzerland/ZH-ETHZ-390394/2020\|EPI_ISL_72 | ···················· | ···················· | ····················· | ···················· | ···················· | ···················· |
| X | 3018 | hCoV-19/Switzerland/AG-ETHZ-340148/2020\|EPI_ISL_72 | ···················· | ···················· | ····················· | ···················· | ···················· | ···················· |
| X | 3019 | hCoV-19/Switzerland/BE-ETHZ-340098/2020\|EPI_ISL_72 | ···················· | ···················· | ····················· | ···················· | ···················· | ···················· |
| X | 3020 | hCoV-19/Switzerland/GR-ETHZ-390411/2020\|EPI_ISL_72 | ···················· | ···················· | ····················· | ···················· | ···················· | ···················· |
| X | 3021 | hCoV-19/Switzerland/ZH-ETHZ-340160/2020\|EPI_ISL_72 | ···················· | ···················· | ····················· | ···················· | ···················· | ···················· |
| X | 3022 | hCoV-19/Switzerland/VS-ETHZ-390403/2020\|EPI_ISL_72 | ···················· | ···················· | ····················· | ···················· | ···················· | ···················· |
| X | 3023 | hCoV-19/Switzerland/AG-ETHZ-340116/2020\|EPI_ISL_72 | ···················· | ···················· | ····················· | ···················· | ···················· | ···················· |
| X | 3024 | hCoV-19/Switzerland/AG-ETHZ-340132/2020\|EPI_ISL_72 | ···················· | ···················· | ····················· | ···················· | ···················· | ···················· |
| X | 3025 | hCoV-19/Switzerland/AG-ETHZ-340135/2020\|EPI_ISL_72 | ···················· | ···················· | ····················· | ···················· | ···················· | ···················· |
| X | 3026 | hCoV-19/Switzerland/AG-ETHZ-340137/2020\|EPI_ISL_72 | ···················· | ···················· | ····················· | ···················· | ···················· | ···················· |
| X | 3027 | hCoV-19/Switzerland/AG-ETHZ-340138/2020\|EPI_ISL_72 | ···················· | ···················· | ····················· | ···················· | ···················· | ···················· |
| X | 3028 | hCoV-19/Switzerland/AG-ETHZ-340139/2020\|EPI_ISL_72 | ···················· | ···················· | ····················· | ···················· | ···················· | ···················· |
| X | 3029 | hCoV-19/Switzerland/AG-ETHZ-340143/2020\|EPI_ISL_72 | ···················· | ···················· | ····················· | ···················· | ···················· | ···················· |
| X | 3030 | hCoV-19/Switzerland/AG-ETHZ-340149/2020\|EPI_ISL_72 | ···················· | ···················· | ····················· | ···················· | ···················· | ···················· |
| X | 3031 | hCoV-19/Switzerland/AG-ETHZ-380234/2020\|EPI_ISL_72 | ···················· | ···················· | ····················· | ···················· | ···················· | ···················· |
| X | 3032 | hCoV-19/Switzerland/AG-ETHZ-380240/2020\|EPI_ISL_72 | ···················· | ···················· | ····················· | ···················· | ···················· | ···················· |
| X | 3033 | hCoV-19/Switzerland/AG-ETHZ-380242/2020\|EPI_ISL_72 | ···················· | ···················· | ····················· | ···················· | ···················· | ···················· |
| X | 3034 | hCoV-19/Switzerland/AG-ETHZ-380246/2020\|EPI_ISL_72 | ···················· | ···················· | ····················· | ···················· | ······K············· | ···················· |
| X | 3035 | hCoV-19/Switzerland/AG-ETHZ-380247/2020\|EPI_ISL_72 | ···················· | ···················· | ····················· | ···················· | ···················· | ···················· |
| X | 3036 | hCoV-19/Switzerland/AG-ETHZ-380282/2020\|EPI_ISL_72 | ···········K········ | ···················· | ····················· | ···················· | ···················· | ···················· |
| X | 3037 | hCoV-19/Switzerland/AG-ETHZ-380287/2020\|EPI_ISL_72 | ···················· | ···················· | ····················· | ···················· | ···················· | ···················· |
| X | 3038 | hCoV-19/Switzerland/AG-ETHZ-380291/2020\|EPI_ISL_72 | ···················· | ·········A·········· | ····················· | ···················· | ···················· | ···················· |
| X | 3039 | hCoV-19/Switzerland/AG-ETHZ-380292/2020\|EPI_ISL_72 | ···················· | ···················· | ····················· | ···················· | ···················· | ···················· |
| X | 3040 | hCoV-19/Switzerland/AG-ETHZ-380293/2020\|EPI_ISL_72 | ···················· | ···················· | ····················· | ···················· | ···················· | ···················· |
| X | 3041 | hCoV-19/Switzerland/AG-ETHZ-380296/2020\|EPI_ISL_72 | ···················· | ···················· | ····················· | ···················· | ···················· | ···················· |
| X | 3042 | hCoV-19/Switzerland/AG-ETHZ-380299/2020\|EPI_ISL_72 | ···················· | ···················· | ····················· | ···················· | ···················· | ···················· |
| X | 3043 | hCoV-19/Switzerland/AG-ETHZ-380300/2020\|EPI_ISL_72 | ···················· | ·········R·········· | ····················· | ···················· | ···················· | ···················· |
| X | 3044 | hCoV-19/Switzerland/AG-ETHZ-380322/2020\|EPI_ISL_72 | ···················· | ···················· | ····················· | ···················· | ···················· | ···················· |
| X | 3045 | hCoV-19/Switzerland/AG-ETHZ-380323/2020\|EPI_ISL_72 | ···················· | ···················· | ····················· | ···················· | ···················· | ···················· |
| X | 3046 | hCoV-19/Switzerland/AG-ETHZ-380324/2020\|EPI_ISL_72 | ···················· | ···················· | ····················· | ···················· | ···················· | ···················· |
| X | 3047 | hCoV-19/Switzerland/AG-ETHZ-380326/2020\|EPI_ISL_72 | ···················· | ···················· | ····················· | ···················· | ···················· | ···················· |
| X | 3048 | hCoV-19/Switzerland/AG-ETHZ-380330/2020\|EPI_ISL_72 | ···················· | ···················· | ····················· | ···················· | ···················· | ···················· |
| X | 3049 | hCoV-19/Switzerland/AG-ETHZ-380337/2020\|EPI_ISL_72 | ···················· | ···················· | ····················· | ···················· | ···················· | ···················· |
| X | 3050 | hCoV-19/Switzerland/AG-ETHZ-380338/2020\|EPI_ISL_72 | ···················· | ···················· | ····················· | ···················· | ···················· | ···················· |
| X | 3051 | hCoV-19/Switzerland/AG-ETHZ-390230/2020\|EPI_ISL_72 | ···················· | ···················· | ····················· | ···················· | ···················· | ···················· |
| X | 3052 | hCoV-19/Switzerland/AG-ETHZ-390231/2020\|EPI_ISL_72 | ···················· | ···················· | ····················· | ···················· | ···················· | ···················· |
| X | 3053 | hCoV-19/Switzerland/AG-ETHZ-390232/2020\|EPI_ISL_72 | ···················· | ···················· | ····················· | ···················· | ···················· | ···················· |
| X | 3054 | hCoV-19/Switzerland/AR-ETHZ-340128/2020\|EPI_ISL_72 | ···················· | ···················· | ····················· | ···················· | ···················· | ···················· |
| X | 3055 | hCoV-19/Switzerland/AR-ETHZ-340152/2020\|EPI_ISL_72 | ···················· | ···················· | ····················· | ···················· | ···················· | ···················· |

| | | | | | | | |
|---|---|---|---|---|---|---|---|
| X | 3056 hCoV-19/Switzerland/AR-ETHZ-340164/2020\|EPI_ISL_72 | ···················· | ···················· | ····················· | ···················· | ···················· | ···················· |
| X | 3057 hCoV-19/Switzerland/AR-ETHZ-390219/2020\|EPI_ISL_72 | ···················· | ···················· | ····················· | ···················· | ···················· | ···················· |
| X | 3058 hCoV-19/Switzerland/AR-ETHZ-390378/2020\|EPI_ISL_72 | ···················· | ···················· | ····················· | ···················· | ···················· | ···················· |
| X | 3059 hCoV-19/Switzerland/BE-ETHZ-340091/2020\|EPI_ISL_72 | ···················· | ···················· | ····················· | ···················· | ···················· | ···················· |
| X | 3060 hCoV-19/Switzerland/BE-ETHZ-340092/2020\|EPI_ISL_72 | ···················· | ···················· | ····················· | ···················· | ···················· | ···················· |
| X | 3061 hCoV-19/Switzerland/BE-ETHZ-340093/2020\|EPI_ISL_72 | ···················· | ···················· | ····················· | ···················· | ···················· | ···················· |
| X | 3062 hCoV-19/Switzerland/BE-ETHZ-340095/2020\|EPI_ISL_72 | ···················· | ···················· | ····················· | ···················· | ···················· | ···················· |
| X | 3063 hCoV-19/Switzerland/BE-ETHZ-340097/2020\|EPI_ISL_72 | ···················· | ···················· | ····················· | ···················· | ···················· | ···················· |
| X | 3064 hCoV-19/Switzerland/BE-ETHZ-340100/2020\|EPI_ISL_72 | ···················· | ···················· | ····················· | ···················· | ···················· | ···················· |
| X | 3065 hCoV-19/Switzerland/BE-ETHZ-340104/2020\|EPI_ISL_72 | ···················· | ···················· | ····················· | ···················· | ···················· | ···················· |
| X | 3066 hCoV-19/Switzerland/BE-ETHZ-340110/2020\|EPI_ISL_72 | ···················· | ···················· | ····················· | ···················· | ···················· | ···················· |
| X | 3067 hCoV-19/Switzerland/BE-ETHZ-340115/2020\|EPI_ISL_72 | ···················· | ···················· | ····················· | ···················· | ···················· | ···················· |
| X | 3068 hCoV-19/Switzerland/BE-ETHZ-340154/2020\|EPI_ISL_72 | ···················· | ···················· | ····················· | ···················· | ···················· | ···················· |
| X | 3069 hCoV-19/Switzerland/BE-ETHZ-340155/2020\|EPI_ISL_72 | ···················· | ···················· | ····················· | ···················· | ······K············· | ···················· |
| X | 3070 hCoV-19/Switzerland/BE-ETHZ-340156/2020\|EPI_ISL_72 | ···················· | ···················· | ····················· | ···················· | ···················· | ···················· |
| X | 3071 hCoV-19/Switzerland/BE-ETHZ-340157/2020\|EPI_ISL_72 | ···················· | ···················· | ····················· | ···················· | ···················· | ···················· |
| X | 3072 hCoV-19/Switzerland/BE-ETHZ-340159/2020\|EPI_ISL_72 | ···················· | ···················· | ····················· | ···················· | ···················· | ···················· |
| X | 3073 hCoV-19/Denmark/DCGC-14814/2020\|EPI_ISL_712002\|202 | ···················· | ···················· | ····················· | ···················· | ···················· | ···················· |
| X | 3074 hCoV-19/Denmark/DCGC-14816/2020\|EPI_ISL_712004\|202 | ···················· | ···················· | ····················· | ···················· | ···················· | ···················· |
| X | 3075 hCoV-19/Denmark/DCGC-14817/2020\|EPI_ISL_712005\|202 | ···················· | ···················· | ····················· | ···················· | ···················· | ···················· |
| X | 3076 hCoV-19/Denmark/DCGC-14818/2020\|EPI_ISL_712006\|202 | ···················· | ···················· | ····················· | ···················· | ···················· | ···················· |
| X | 3077 hCoV-19/Denmark/DCGC-14819/2020\|EPI_ISL_712007\|202 | ···················· | ···················· | ····················· | ···················· | ···················· | ···················· |
| X | 3078 hCoV-19/Denmark/DCGC-14820/2020\|EPI_ISL_712008\|202 | ···················· | ···················· | ····················· | ···················· | ···················· | ···················· |
| X | 3079 hCoV-19/Denmark/DCGC-14821/2020\|EPI_ISL_712009\|202 | ···················· | ···················· | ····················· | ···················· | ···················· | ···················· |
| X | 3080 hCoV-19/Denmark/DCGC-15118/2020\|EPI_ISL_713319\|202 | ···················· | ···················· | ····················· | ···················· | ···················· | ···················· |
| X | 3081 hCoV-19/Denmark/DCGC-15159/2020\|EPI_ISL_713320\|202 | ···················· | ···················· | ····················· | ···················· | ···················· | ···················· |
| X | 3082 hCoV-19/Denmark/DCGC-15250/2020\|EPI_ISL_713321\|202 | ···················· | ···················· | ····················· | ···················· | ···················· | ···················· |
| X | 3083 hCoV-19/Denmark/DCGC-15470/2020\|EPI_ISL_713322\|202 | ···················· | ···················· | ····················· | ···················· | ···················· | ···················· |
| X | 3084 hCoV-19/Denmark/DCGC-15595/2020\|EPI_ISL_713323\|202 | ···················· | ···················· | ····················· | ···················· | ···················· | ···················· |
| X | 3085 hCoV-19/Denmark/DCGC-15652/2020\|EPI_ISL_713324\|202 | ···················· | ···················· | ····················· | ···················· | ···················· | ···················· |
| X | 3086 hCoV-19/Denmark/DCGC-15692/2020\|EPI_ISL_713325\|202 | ···················· | ···················· | ····················· | ···················· | ···················· | ···················· |
| X | 3087 hCoV-19/Denmark/DCGC-15699/2020\|EPI_ISL_713326\|202 | ···················· | ···················· | ····················· | ···················· | ···················· | ···················· |
| X | 3088 hCoV-19/Denmark/DCGC-15730/2020\|EPI_ISL_713327\|202 | ···················· | ···················· | ····················· | ···················· | ···················· | ···················· |
| X | 3089 hCoV-19/Denmark/DCGC-15883/2020\|EPI_ISL_713328\|202 | ···················· | ···················· | ····················· | ···················· | ···················· | ···················· |
| X | 3090 hCoV-19/Denmark/DCGC-14938/2020\|EPI_ISL_713334\|202 | ···················· | ···················· | ····················· | ···················· | ···················· | ···················· |
| X | 3091 hCoV-19/Denmark/DCGC-14957/2020\|EPI_ISL_713335\|202 | ···················· | ···················· | ····················· | ···················· | ···················· | ···················· |
| X | 3092 hCoV-19/Denmark/DCGC-14989/2020\|EPI_ISL_713336\|202 | ···················· | ···················· | ····················· | ···················· | ···················· | ···················· |
| X | 3093 hCoV-19/Denmark/DCGC-14991/2020\|EPI_ISL_713337\|202 | ···················· | ···················· | ····················· | ···················· | ···················· | ···················· |
| X | 3094 hCoV-19/Denmark/DCGC-15004/2020\|EPI_ISL_713338\|202 | ···················· | ···················· | ····················· | ···················· | ···················· | ···················· |
| X | 3095 hCoV-19/Denmark/DCGC-15031/2020\|EPI_ISL_713339\|202 | ···················· | ···················· | ····················· | ···················· | ···················· | ···················· |
| X | 3096 hCoV-19/Denmark/DCGC-16135/2020\|EPI_ISL_714600\|202 | ···················· | ···················· | ····················· | ···················· | ···················· | ···················· |
| X | 3097 hCoV-19/Denmark/DCGC-16143/2020\|EPI_ISL_714601\|202 | ···················· | ···················· | ····················· | ···················· | ···················· | ···················· |
| X | 3098 hCoV-19/Denmark/DCGC-16144/2020\|EPI_ISL_714602\|202 | ···················· | ···················· | ····················· | ···················· | ···················· | ···················· |
| X | 3099 hCoV-19/Denmark/DCGC-16146/2020\|EPI_ISL_714603\|202 | ···················· | ···················· | ····················· | ···················· | ···················· | ···················· |
| X | 3100 hCoV-19/Denmark/DCGC-16147/2020\|EPI_ISL_714604\|202 | ···················· | ···················· | ····················· | ···················· | ···················· | ···················· |
| X | 3101 hCoV-19/Denmark/DCGC-16149/2020\|EPI_ISL_714605\|202 | ···················· | ···················· | ····················· | ···················· | ···················· | ···················· |
| X | 3102 hCoV-19/Denmark/DCGC-16151/2020\|EPI_ISL_714606\|202 | ···················· | ···················· | ····················· | ···················· | ···················· | ···················· |
| X | 3103 hCoV-19/Denmark/DCGC-16152/2020\|EPI_ISL_714607\|202 | ···················· | ···················· | ····················· | ···················· | ···················· | ···················· |
| X | 3104 hCoV-19/Denmark/DCGC-16153/2020\|EPI_ISL_714608\|202 | ···················· | ···················· | ····················· | ···················· | ···················· | ···················· |
| X | 3105 hCoV-19/Denmark/DCGC-16154/2020\|EPI_ISL_714609\|202 | ···················· | ···················· | ····················· | ···················· | ···················· | ···················· |
| X | 3106 hCoV-19/Denmark/DCGC-16155/2020\|EPI_ISL_714610\|202 | ···················· | ···················· | ····················· | ···················· | ···················· | ···················· |
| X | 3107 hCoV-19/Denmark/DCGC-16157/2020\|EPI_ISL_714611\|202 | ···················· | ···················· | ····················· | ···················· | ···················· | ···················· |
| X | 3108 hCoV-19/Denmark/DCGC-16158/2020\|EPI_ISL_714612\|202 | ···················· | ···················· | ····················· | ···················· | ···················· | ···················· |
| X | 3109 hCoV-19/Denmark/DCGC-16161/2020\|EPI_ISL_714613\|202 | ···················· | ···················· | ····················· | ···················· | ···················· | ···················· |
| X | 3110 hCoV-19/Denmark/DCGC-16162/2020\|EPI_ISL_714614\|202 | ···················· | ··········A········· | ····················· | ···················· | ···················· | ···················· |
| X | 3111 hCoV-19/Denmark/DCGC-16163/2020\|EPI_ISL_714615\|202 | ···················· | ···················· | ····················· | ···················· | ···················· | ···················· |
| X | 3112 hCoV-19/Denmark/DCGC-16164/2020\|EPI_ISL_714616\|202 | ···················· | ···················· | ····················· | ···················· | ···················· | ···················· |
| X | 3113 hCoV-19/Denmark/DCGC-16166/2020\|EPI_ISL_714617\|202 | ···················· | ···················· | ····················· | ···················· | ···················· | ···················· |
| X | 3114 hCoV-19/Denmark/DCGC-16168/2020\|EPI_ISL_714618\|202 | ···················· | ···················· | ····················· | ···················· | ······T············· | ···················· |
| X | 3115 hCoV-19/Denmark/DCGC-16170/2020\|EPI_ISL_714619\|202 | ···················· | ···················· | ····················· | ···················· | ···················· | ···················· |
| X | 3116 hCoV-19/Denmark/DCGC-16173/2020\|EPI_ISL_714620\|202 | ···················· | ···················· | ····················· | ···················· | ···················· | ···················· |
| X | 3117 hCoV-19/Denmark/DCGC-16175/2020\|EPI_ISL_714621\|202 | ···················· | ···················· | ····················· | ···················· | ···················· | ···················· |
| X | 3118 hCoV-19/Denmark/DCGC-16177/2020\|EPI_ISL_714622\|202 | ···················· | ···················· | ····················· | ···················· | ···················· | ···················· |
| X | 3119 hCoV-19/Denmark/DCGC-16178/2020\|EPI_ISL_714623\|202 | ···················· | ·A·················· | ····················· | ···················· | ···················· | ···A················ |
| X | 3120 hCoV-19/Denmark/DCGC-16179/2020\|EPI_ISL_714624\|202 | ···················· | ···················· | ····················· | ···················· | ···················· | ···················· |
| X | 3121 hCoV-19/Denmark/DCGC-16184/2020\|EPI_ISL_714625\|202 | ···················· | ···················· | ····················· | ···················· | ···················· | ···················· |
| X | 3122 hCoV-19/Denmark/DCGC-16185/2020\|EPI_ISL_714626\|202 | ···················· | ···················· | ····················· | ···················· | ···················· | ···················· |
| X | 3123 hCoV-19/Denmark/DCGC-16188/2020\|EPI_ISL_714627\|202 | ···················· | ···················· | ····················· | ···················· | ···················· | ···················· |
| X | 3124 hCoV-19/Denmark/DCGC-16226/2020\|EPI_ISL_714650\|202 | ···················· | ···················· | ····················· | ···················· | ···················· | ···················· |
| X | 3125 hCoV-19/Denmark/DCGC-16227/2020\|EPI_ISL_714651\|202 | ···················· | ···················· | ····················· | ···················· | ···················· | ···················· |
| X | 3126 hCoV-19/Denmark/DCGC-16228/2020\|EPI_ISL_714652\|202 | ···················· | ····A··············· | ····················· | ···················· | ···················· | ···················· |
| X | 3127 hCoV-19/Denmark/DCGC-16229/2020\|EPI_ISL_714653\|202 | ···················· | ···················· | ····················· | ···················· | ···················· | ···················· |
| X | 3128 hCoV-19/Denmark/DCGC-16232/2020\|EPI_ISL_714654\|202 | ···················· | ···················· | ····················· | ···················· | ···················· | ···················· |
| X | 3129 hCoV-19/Denmark/DCGC-16234/2020\|EPI_ISL_714655\|202 | ···················· | ···················· | ····················· | ···················· | ···················· | ···················· |
| X | 3130 hCoV-19/Denmark/DCGC-16235/2020\|EPI_ISL_714656\|202 | ···················· | ···················· | ····················· | ···················· | ···················· | ···················· |
| X | 3131 hCoV-19/Denmark/DCGC-16236/2020\|EPI_ISL_714657\|202 | ···················· | ···················· | ····················· | ···················· | ···················· | ···················· |
| X | 3132 hCoV-19/Denmark/DCGC-16238/2020\|EPI_ISL_714658\|202 | ···················· | ···················· | ····················· | ···················· | ······T············· | ···················· |
| X | 3133 hCoV-19/Denmark/DCGC-16239/2020\|EPI_ISL_714659\|202 | ···················· | ···················· | ····················· | ···················· | ···················· | ···················· |
| X | 3134 hCoV-19/Denmark/DCGC-16248/2020\|EPI_ISL_714664\|202 | ···················· | ·A·················· | ····················· | ···················· | ···················· | ···················· |
| X | 3135 hCoV-19/Denmark/DCGC-16249/2020\|EPI_ISL_714665\|202 | ···················· | ···················· | ····················· | ···················· | ···················· | ···················· |

| | | | | | | | | |
|---|---|---|---|---|---|---|---|---|
| X | 3136 | hCoV-19/Denmark/DCGC-16253/2020\|EPI_ISL_714666\|202 | .................... | .................... | .................... | .................... | .................... | .................... |
| X | 3137 | hCoV-19/Denmark/DCGC-16254/2020\|EPI_ISL_714667\|202 | .................... | .................... | .................... | .................... | .................... | .................... |
| X | 3138 | hCoV-19/Denmark/DCGC-16256/2020\|EPI_ISL_714668\|202 | .................... | .................... | .................... | .................... | .................... | .................... |
| X | 3139 | hCoV-19/Denmark/DCGC-16257/2020\|EPI_ISL_714669\|202 | .................... | .................... | .................... | .................... | .................... | .................... |
| X | 3140 | hCoV-19/Malaysia/UNIMAS-0729/2020\|EPI_ISL_718132\|2 | .................... | .................... | .................... | .................... | .................... | .................... |
| X | 3141 | hCoV-19/Malaysia/UNIMAS-M3851/2020\|EPI_ISL_718133\| | .................... | .................... | .................... | .................... | .................... | .................... |
| X | 3142 | hCoV-19/Malaysia/UNIMAS-C122/2020\|EPI_ISL_718134\|2 | .................... | .................... | .................... | .................... | .................... | .................... |
| X | 3143 | hCoV-19/Malaysia/UNIMAS-C121/2020\|EPI_ISL_718135\|2 | .................... | .................... | .................... | .................... | .................... | .................... |
| X | 3144 | hCoV-19/Malaysia/UNIMAS-0803/2020\|EPI_ISL_718136\|2 | .................... | .................... | .................... | .................... | .................... | .................... |
| X | 3145 | hCoV-19/Malaysia/UNIMAS-0694/2020\|EPI_ISL_718137\|2 | .................... | .................... | .................... | .................... | .................... | .................... |
| X | 3146 | hCoV-19/Malaysia/UNIMAS-0298/2020\|EPI_ISL_718138\|2 | .................... | .................... | .................... | .................... | .................... | .................... |
| X | 3147 | hCoV-19/Malaysia/UNIMAS-3325/2020\|EPI_ISL_718139\|2 | .................... | .................... | .................... | .................... | .................... | .................... |
| X | 3148 | hCoV-19/Malaysia/UNIMAS-0179/2020\|EPI_ISL_718140\|2 | .................... | .................... | .................... | .................... | .................... | .................... |
| X | 3149 | hCoV-19/Malaysia/UNIMAS-2615/2020\|EPI_ISL_718141\|2 | .................... | .................... | .................... | .................... | .................... | .................... |
| X | 3150 | hCoV-19/Malaysia/UNIMAS-0212/2020\|EPI_ISL_718142\|2 | .................... | .................... | .................... | .................... | .................... | .................... |
| X | 3151 | hCoV-19/Malaysia/UNIMAS-14960/2020\|EPI_ISL_718143\| | .................... | .................... | .................... | .................... | .................... | .................... |
| X | 3152 | hCoV-19/Malaysia/UNIMAS-0304/2020\|EPI_ISL_718144\|2 | .................... | .................... | .................... | .................... | .................... | .................... |
| X | 3153 | hCoV-19/Malaysia/UNIMAS-M0710/2020\|EPI_ISL_718145\| | .................... | .................... | .................... | .................... | .................... | .................... |
| X | 3154 | hCoV-19/Malaysia/UNIMAS-11942/2020\|EPI_ISL_718146\| | .................... | .................... | .................... | .................... | .................... | .................... |
| X | 3155 | hCoV-19/Malaysia/UNIMAS-0358/2020\|EPI_ISL_718148\|2 | .................... | .................... | .................... | .................... | .................... | .................... |
| X | 3156 | hCoV-19/Malaysia/UNIMAS-1020/2020\|EPI_ISL_718149\|2 | .................... | .................... | .................... | .................... | .................... | .................... |
| X | 3157 | hCoV-19/Malaysia/UNIMAS-0172/2020\|EPI_ISL_718150\|2 | .................... | .................... | .................... | .................... | .................... | .................... |
| X | 3158 | hCoV-19/Malaysia/UNIMAS-1741/2020\|EPI_ISL_718151\|2 | .................... | .................... | .................... | .................... | .................... | .................... |
| X | 3159 | hCoV-19/Malaysia/UNIMAS-0136/2020\|EPI_ISL_718152\|2 | .................... | .................... | .................... | .................... | .................... | .................... |
| X | 3160 | hCoV-19/Malaysia/UNIMAS-1589/2020\|EPI_ISL_718153\|2 | .................... | .................... | .................... | .................... | .................... | .................... |
| X | 3161 | hCoV-19/Malaysia/UNIMAS-M3002/2020\|EPI_ISL_718154\| | .................... | .................... | .................... | .................... | .................... | .................... |
| X | 3162 | hCoV-19/Malaysia/UNIMAS-2304/2020\|EPI_ISL_718155\|2 | .................... | .................... | .................... | .................... | .................... | .................... |
| X | 3163 | hCoV-19/Malaysia/UNIMAS-C012N/2020\|EPI_ISL_718156\| | .................... | .................... | .................... | .................... | .................... | .................... |
| X | 3164 | hCoV-19/Malaysia/UNIMAS-0243/2020\|EPI_ISL_718157\|2 | .................... | .................... | .................... | .................... | .................... | .................... |
| X | 3165 | hCoV-19/Malaysia/UNIMAS-4248R/2020\|EPI_ISL_718158\| | .................... | .................... | .................... | .................... | .................... | .................... |
| X | 3166 | hCoV-19/Malaysia/UNIMAS-M1884/2020\|EPI_ISL_718159\| | .................... | .................... | .................... | .................... | .................... | .................... |
| X | 3167 | hCoV-19/Malaysia/UNIMAS-M0872/2020\|EPI_ISL_718160\| | .................... | .................... | .................... | .................... | .................... | .................... |
| X | 3168 | hCoV-19/Malaysia/UNIMAS-1197/2020\|EPI_ISL_718161\|2 | .................... | .................... | .................... | .................... | .................... | .................... |
| X | 3169 | hCoV-19/Malaysia/UNIMAS-0242/2020\|EPI_ISL_718162\|2 | .................... | .................... | .................... | .................... | .................... | .................... |
| X | 3170 | hCoV-19/Malaysia/UNIMAS-0099/2020\|EPI_ISL_718163\|2 | .................... | .................... | .................... | .................... | .................... | .................... |
| X | 3171 | hCoV-19/Malaysia/UNIMAS-0217/2020\|EPI_ISL_718164\|2 | .................... | .................... | .................... | .................... | .................... | .................... |
| X | 3172 | hCoV-19/Malaysia/UNIMAS-M4061/2020\|EPI_ISL_718165\| | .................... | .................... | .................... | .................... | .................... | .................... |
| X | 3173 | hCoV-19/Malaysia/UNIMAS-M3482/2020\|EPI_ISL_718166\| | .................... | .................... | .................... | .................... | .................... | .................... |
| X | 3174 | hCoV-19/Malaysia/UNIMAS-B066/2020\|EPI_ISL_718167\|2 | .................... | .................... | .................... | .................... | .................... | .................... |
| X | 3175 | hCoV-19/Malaysia/UNIMAS-15695/2020\|EPI_ISL_718168\| | .................... | .................... | .................... | .................... | .................... | .................... |
| X | 3176 | hCoV-19/Malaysia/UNIMAS-15723/2020\|EPI_ISL_718169\| | .................... | .A.................. | .................... | .................... | .................... | .................... |
| X | 3177 | hCoV-19/Malaysia/UNIMAS-15729/2020\|EPI_ISL_718170\| | .................... | .A.................. | .................... | .................... | .................... | .................... |
| X | 3178 | hCoV-19/Malaysia/UNIMAS-15720/2020\|EPI_ISL_718171\| | .................... | .A.................. | .................... | .................... | .................... | .................... |
| X | 3179 | hCoV-19/Malaysia/UNIMAS-15790/2020\|EPI_ISL_718172\| | .................... | .A.................. | .................... | .................... | .................... | .................... |
| X | 3180 | hCoV-19/Malaysia/UNIMAS-15735/2020\|EPI_ISL_718173\| | .................... | .A.................. | .................... | .................... | .................... | .................... |
| X | 3181 | hCoV-19/Malaysia/UNIMAS-B045/2020\|EPI_ISL_718174\|2 | .................... | .A.................. | .................... | .................... | .................... | .................... |
| X | 3182 | hCoV-19/Malaysia/UNIMAS-15732/2020\|EPI_ISL_718175\| | .................... | .A.................. | .................... | .................... | .................... | .................... |
| X | 3183 | hCoV-19/Malaysia/UNIMAS-B074/2020\|EPI_ISL_718176\|2 | .................... | .A.................. | .................... | .................... | .................... | .................... |
| X | 3184 | hCoV-19/Malaysia/UNIMAS-15721/2020\|EPI_ISL_718177\| | .................... | .A.................. | .................... | .................... | .................... | .................... |
| X | 3185 | hCoV-19/Malaysia/UNIMAS-B040/2020\|EPI_ISL_718178\|2 | .................... | .A.................. | .................... | .................... | .................... | .................... |
| X | 3186 | hCoV-19/Malaysia/UNIMAS-15755/2020\|EPI_ISL_718179\| | .................... | .A.................. | .................... | .................... | .................... | .................... |
| X | 3187 | hCoV-19/Malaysia/UNIMAS-15768/2020\|EPI_ISL_718180\| | .................... | .A.................. | .................... | .................... | .................... | .................... |
| X | 3188 | hCoV-19/Malaysia/UNIMAS-15783/2020\|EPI_ISL_718181\| | .................... | .................... | .................... | .................... | .................... | .................... |
| X | 3189 | hCoV-19/Malaysia/UNIMAS-B051/2020\|EPI_ISL_718182\|2 | .................... | .A.................. | .................... | .................... | .................... | .................... |
| X | 3190 | hCoV-19/Malaysia/UNIMAS-15762/2020\|EPI_ISL_718183\| | .................... | .A.................. | .................... | .................... | .................... | .................... |
| X | 3191 | hCoV-19/Malaysia/UNIMAS-15765/2020\|EPI_ISL_718184\| | .................... | .A.................. | .................... | .................... | .................... | .................... |
| X | 3192 | hCoV-19/Malaysia/UNIMAS-B073/2020\|EPI_ISL_718185\|2 | .................... | .A.................. | .................... | .................... | .................... | .................... |
| X | 3193 | hCoV-19/Malaysia/UNIMAS-15764/2020\|EPI_ISL_718186\| | .................... | .A.................. | .................... | .................... | .................... | .................... |
| X | 3194 | hCoV-19/Malaysia/UNIMAS-15780/2020\|EPI_ISL_718187\| | .................... | .A.................. | .................... | .................... | .................... | .................... |
| X | 3195 | hCoV-19/Malaysia/UNIMAS-15745/2020\|EPI_ISL_718188\| | .................... | .A.................. | .................... | .................... | .................... | .................... |
| X | 3196 | hCoV-19/Malaysia/UNIMAS-15763/2020\|EPI_ISL_718189\| | .................... | .A.................. | .................... | .................... | .................... | .................... |
| X | 3197 | hCoV-19/Malaysia/UNIMAS-15758/2020\|EPI_ISL_718190\| | .................... | .A.................. | .................... | .................... | .................... | .................... |
| X | 3198 | hCoV-19/Malaysia/UNIMAS-B057/2020\|EPI_ISL_718191\|2 | .................... | .A.................. | .................... | .................... | .................... | .................... |
| X | 3199 | hCoV-19/Malaysia/UNIMAS-B022/2020\|EPI_ISL_718192\|2 | .................... | .A.................. | .................... | .................... | .................... | .................... |
| X | 3200 | hCoV-19/Malaysia/UNIMAS-15772/2020\|EPI_ISL_718193\| | .................... | .A.................. | .................... | .................... | .................... | .................... |
| X | 3201 | hCoV-19/Malaysia/UNIMAS-15777/2020\|EPI_ISL_718194\| | .................... | .A.................. | .................... | .................... | .................... | .................... |
| X | 3202 | hCoV-19/Malaysia/UNIMAS-15782/2020\|EPI_ISL_718195\| | .................... | .A.................. | .................... | .................... | .................... | .................... |
| X | 3203 | hCoV-19/Malaysia/UNIMAS-15785/2020\|EPI_ISL_718196\| | .................... | .A.................. | .................... | .................... | .................... | .................... |
| X | 3204 | hCoV-19/Malaysia/UNIMAS-B028/2020\|EPI_ISL_718197\|2 | .................... | .A.................. | .................... | .................... | .................... | .................... |
| X | 3205 | hCoV-19/Malaysia/UNIMAS-B037/2020\|EPI_ISL_718198\|2 | .................... | .A.................. | .................... | .................... | .................... | .................... |
| X | 3206 | hCoV-19/Malaysia/UNIMAS-15766/2020\|EPI_ISL_718199\| | .................... | .A.................. | .................... | .................... | .................... | .................... |
| X | 3207 | hCoV-19/England/CAMC-C44189/2020\|EPI_ISL_719445\|20 | .................... | .................... | .................... | .................... | .................... | .................... |
| X | 3208 | hCoV-19/England/CAMC-C4420E/2020\|EPI_ISL_719447\|20 | .................... | .................... | .................... | .................... | .................... | .................... |
| X | 3209 | hCoV-19/England/CAMC-C3FCCC/2020\|EPI_ISL_719454\|20 | .................... | .................... | .................... | .................... | .................... | .................... |
| X | 3210 | hCoV-19/England/CAMC-C44AF7/2020\|EPI_ISL_719455\|20 | .................... | .................... | .................... | .................... | .................... | .................... |
| X | 3211 | hCoV-19/England/CAMC-C43F80/2020\|EPI_ISL_719459\|20 | .................... | .................... | .................... | .................... | .................... | .................... |
| X | 3212 | hCoV-19/England/CAMC-C45829/2020\|EPI_ISL_719460\|20 | .................... | .................... | .................... | .................... | .................... | .................... |
| X | 3213 | hCoV-19/England/CAMC-C4324B/2020\|EPI_ISL_719463\|20 | .................... | .................... | .................... | .................... | .................... | .................... |
| X | 3214 | hCoV-19/England/CAMC-C448A2/2020\|EPI_ISL_719466\|20 | .................... | .................... | .................... | .................... | .................... | .................... |
| X | 3215 | hCoV-19/England/CAMC-C3FEE4/2020\|EPI_ISL_719471\|20 | .................... | .................... | .................... | .................... | .................... | .................... |

| | | | | | | | |
|---|---|---|---|---|---|---|---|
| X | 3216 hCoV-19/England/CAMC-C43551/2020\|EPI_ISL_719472\|20 | ···················· | ···················· | ···················· | ···················· | ···················· | ···················· |
| X | 3217 hCoV-19/England/CAMC-C3F3C7/2020\|EPI_ISL_719473\|20 | ···················· | ···················· | ···················· | ···················· | ···················· | ···················· |
| X | 3218 hCoV-19/England/CAMC-C43560/2020\|EPI_ISL_719478\|20 | ···················· | ···················· | ···················· | ···················· | ···················· | ···················· |
| X | 3219 hCoV-19/England/CAMC-C44875/2020\|EPI_ISL_719481\|20 | ···················· | ···················· | ···················· | ···················· | ···················· | ···················· |
| X | 3220 hCoV-19/England/CAMC-C44338/2020\|EPI_ISL_719482\|20 | ···················· | ···················· | ···················· | ···················· | ···················· | ···················· |
| X | 3221 hCoV-19/England/CAMC-C43542/2020\|EPI_ISL_719484\|20 | ···················· | ···················· | ···················· | ···················· | ······T············· | ···················· |
| X | 3222 hCoV-19/England/CAMC-C43E74/2020\|EPI_ISL_719485\|20 | ···················· | ···················· | ···················· | ···················· | ···················· | ···················· |
| X | 3223 hCoV-19/England/CAMC-C3F39A/2020\|EPI_ISL_719490\|20 | ···················· | ···················· | ···················· | ···················· | ···················· | ···················· |
| X | 3224 hCoV-19/England/MILK-BB105E/2020\|EPI_ISL_720425\|20 | ···················· | ···················· | ···················· | ···················· | ···················· | ···················· |
| X | 3225 hCoV-19/England/MILK-BABAA4/2020\|EPI_ISL_720426\|20 | ···················· | ···················· | ···················· | ···················· | ···················· | ···················· |
| X | 3226 hCoV-19/England/MILK-BB1FF7/2020\|EPI_ISL_720427\|20 | ···················· | ···················· | ···················· | ···················· | ···················· | ···················· |
| X | 3227 hCoV-19/England/MILK-BAC5DF/2020\|EPI_ISL_720428\|20 | ···················· | ···················· | ···················· | ···················· | ···················· | ···················· |
| X | 3228 hCoV-19/England/MILK-BB0DC1/2020\|EPI_ISL_720429\|20 | ···················· | ···················· | ···················· | ···················· | ···················· | ···················· |
| X | 3229 hCoV-19/England/MILK-BAC399/2020\|EPI_ISL_720430\|20 | ···················· | ···················· | ···················· | ···················· | ···················· | ···················· |
| X | 3230 hCoV-19/England/MILK-BB0BB8/2020\|EPI_ISL_720431\|20 | ···················· | ···················· | ···················· | ···················· | ······T············· | ···················· |
| X | 3231 hCoV-19/England/MILK-BB1FE8/2020\|EPI_ISL_720432\|20 | ···················· | ···················· | ···················· | ···················· | ···················· | ···················· |
| X | 3232 hCoV-19/England/MILK-BB0031/2020\|EPI_ISL_720433\|20 | ···················· | ···················· | ···················· | ···················· | ···················· | ···················· |
| X | 3233 hCoV-19/England/MILK-BB0F52/2020\|EPI_ISL_720434\|20 | ···················· | ···················· | ···················· | ···················· | ···················· | ···················· |
| X | 3234 hCoV-19/England/MILK-BB0848/2020\|EPI_ISL_720435\|20 | ···················· | ···················· | ···················· | ···················· | ······T············· | ···················· |
| X | 3235 hCoV-19/England/MILK-BB1CE1/2020\|EPI_ISL_720436\|20 | ···················· | ···················· | ···················· | ···················· | ···················· | ···················· |
| X | 3236 hCoV-19/England/MILK-BAC5A2/2020\|EPI_ISL_720438\|20 | ···················· | ···················· | ···················· | ···················· | ···················· | ···················· |
| X | 3237 hCoV-19/England/MILK-BB0E55/2020\|EPI_ISL_720440\|20 | ···················· | ···················· | ···················· | ···················· | ···················· | ···················· |
| X | 3238 hCoV-19/England/MILK-BAC593/2020\|EPI_ISL_720441\|20 | ···················· | ···················· | ···················· | ···················· | ···················· | ···················· |
| X | 3239 hCoV-19/England/MILK-BB1D1B/2020\|EPI_ISL_720442\|20 | ···················· | ···················· | ···················· | ···················· | ···················· | ···················· |
| X | 3240 hCoV-19/England/MILK-BABB1A/2020\|EPI_ISL_720443\|20 | ···················· | ···················· | ···················· | ···················· | ···················· | ···················· |
| X | 3241 hCoV-19/England/MILK-BB1AD8/2020\|EPI_ISL_720444\|20 | ···················· | ···················· | ···················· | ···················· | ···················· | ···················· |
| X | 3242 hCoV-19/England/MILK-BB07F0/2020\|EPI_ISL_720447\|20 | ···················· | ···················· | ···················· | ···················· | ···················· | ···················· |
| X | 3243 hCoV-19/England/MILK-BB03A1/2020\|EPI_ISL_720448\|20 | ···················· | ···················· | ···················· | ···················· | ···················· | ···················· |
| X | 3244 hCoV-19/England/MILK-BAC5EE/2020\|EPI_ISL_720449\|20 | ···················· | ···················· | ···················· | ···················· | ···················· | ···················· |
| X | 3245 hCoV-19/England/MILK-BAB8D7/2020\|EPI_ISL_720451\|20 | ···················· | ···················· | ···················· | ···················· | ···················· | ···················· |
| X | 3246 hCoV-19/England/MILK-BAB910/2020\|EPI_ISL_720454\|20 | ···················· | ···················· | ···················· | ···················· | ···················· | ···················· |
| X | 3247 hCoV-19/England/MILK-BB1ABA/2020\|EPI_ISL_720455\|20 | ···················· | ···················· | ···················· | ···················· | ···················· | ···················· |
| X | 3248 hCoV-19/England/MILK-BABAC2/2020\|EPI_ISL_720456\|20 | ···················· | ···················· | ···················· | ···················· | ···················· | ···················· |
| X | 3249 hCoV-19/England/MILK-BABF69/2020\|EPI_ISL_720457\|20 | ···················· | ···················· | ···················· | ···················· | ···················· | ···················· |
| X | 3250 hCoV-19/England/MILK-BB0ACA/2020\|EPI_ISL_720458\|20 | ···················· | ·A·················· | ···················· | ···················· | ···················· | ··A················· |
| X | 3251 hCoV-19/England/MILK-BAFFFC/2020\|EPI_ISL_720459\|20 | ···················· | ···················· | ···················· | ···················· | ···················· | ···················· |
| X | 3252 hCoV-19/England/MILK-BB113D/2020\|EPI_ISL_720460\|20 | ···················· | ···················· | ···················· | ···················· | ···················· | ···················· |
| X | 3253 hCoV-19/England/MILK-BB115B/2020\|EPI_ISL_720463\|20 | ···················· | ···················· | ···················· | ···················· | ···················· | ···················· |
| X | 3254 hCoV-19/England/MILK-BB19EA/2020\|EPI_ISL_720464\|20 | ···················· | ···················· | ···················· | ···················· | ···················· | ···················· |
| X | 3255 hCoV-19/England/MILK-BB21A5/2020\|EPI_ISL_720465\|20 | ···················· | ···················· | ···················· | ···················· | ···················· | ···················· |
| X | 3256 hCoV-19/England/MILK-BAC42D/2020\|EPI_ISL_720468\|20 | ···················· | ···················· | ···················· | ···················· | ···················· | ···················· |
| X | 3257 hCoV-19/England/MILK-BB1DA2/2020\|EPI_ISL_720470\|20 | ···················· | ···················· | ···················· | ···················· | ···················· | ··A················· |
| X | 3258 hCoV-19/England/MILK-BB0B21/2020\|EPI_ISL_720471\|20 | ···················· | ···················· | ···················· | ···················· | ···················· | ···················· |
| X | 3259 hCoV-19/England/MILK-BAFC13/2020\|EPI_ISL_720472\|20 | ···················· | ···················· | ···················· | ···················· | ···················· | ···················· |
| X | 3260 hCoV-19/England/MILK-BB22B1/2020\|EPI_ISL_720473\|20 | ···················· | ···················· | ···················· | ···················· | ···················· | ···················· |
| X | 3261 hCoV-19/England/MILK-BABA95/2020\|EPI_ISL_720474\|20 | ···················· | ···················· | ···················· | ···················· | ···················· | ···················· |
| X | 3262 hCoV-19/England/MILK-BAFDF2/2020\|EPI_ISL_720475\|20 | ···················· | ···················· | ···················· | ···················· | ···················· | ···················· |
| X | 3263 hCoV-19/England/MILK-BABB0B/2020\|EPI_ISL_720476\|20 | ···················· | ···················· | ···················· | ···················· | ···················· | ···················· |
| X | 3264 hCoV-19/England/MILK-BB19F9/2020\|EPI_ISL_720477\|20 | ···················· | ···················· | ···················· | ···················· | ···················· | ···················· |
| X | 3265 hCoV-19/England/MILK-BB210F/2020\|EPI_ISL_720479\|20 | ···················· | ···················· | ···················· | ···················· | ···················· | ···················· |
| X | 3266 hCoV-19/England/MILK-BB0013/2020\|EPI_ISL_720481\|20 | ···················· | ···················· | ···················· | ···················· | ···················· | ···················· |
| X | 3267 hCoV-19/England/MILK-BB0BD6/2020\|EPI_ISL_720482\|20 | ···················· | ···················· | ···················· | ···················· | ···················· | ···················· |
| X | 3268 hCoV-19/England/MILK-BB0BE5/2020\|EPI_ISL_720483\|20 | ···················· | ···················· | ···················· | ···················· | ···················· | ···················· |
| X | 3269 hCoV-19/England/MILK-BB0E0A/2020\|EPI_ISL_720484\|20 | ···················· | ···················· | ···················· | ···················· | ···················· | ···················· |
| X | 3270 hCoV-19/England/MILK-BABF78/2020\|EPI_ISL_720485\|20 | ···················· | ···················· | ···················· | ···················· | ···················· | ···················· |
| X | 3271 hCoV-19/England/MILK-BB0C88/2020\|EPI_ISL_720486\|20 | ···················· | ···················· | ···················· | ···················· | ···················· | ···················· |
| X | 3272 hCoV-19/England/MILK-BABF87/2020\|EPI_ISL_720487\|20 | ···················· | ···················· | ···················· | ···················· | ···················· | ···················· |
| X | 3273 hCoV-19/England/MILK-BB0CC4/2020\|EPI_ISL_720489\|20 | ···················· | ···················· | ···················· | ···················· | ···················· | ···················· |
| X | 3274 hCoV-19/England/MILK-BB06D5/2020\|EPI_ISL_720490\|20 | ···················· | ···················· | ···················· | ···················· | ···················· | ···················· |
| X | 3275 hCoV-19/England/MILK-BAC38A/2020\|EPI_ISL_720491\|20 | ···················· | ···················· | ···················· | ···················· | ······T············· | ···················· |
| X | 3276 hCoV-19/England/MILK-BB2354/2020\|EPI_ISL_720492\|20 | ···················· | ···················· | ···················· | ···················· | ···················· | ···················· |
| X | 3277 hCoV-19/England/MILK-BB1D0C/2020\|EPI_ISL_720493\|20 | ···················· | ···················· | ···················· | ···················· | ···················· | ···················· |
| X | 3278 hCoV-19/England/MILK-BB0884/2020\|EPI_ISL_720495\|20 | ···················· | ···················· | ···················· | ···················· | ···················· | ···················· |
| X | 3279 hCoV-19/England/MILK-BB007D/2020\|EPI_ISL_720496\|20 | ···················· | ···················· | ···················· | ···················· | ···················· | ···················· |
| X | 3280 hCoV-19/England/MILK-BB0C4C/2020\|EPI_ISL_720498\|20 | ···················· | ···················· | ···················· | ···················· | ···················· | ···················· |
| X | 3281 hCoV-19/England/MILK-BB07D2/2020\|EPI_ISL_720499\|20 | ···················· | ···················· | ···················· | ···················· | ···················· | ···················· |
| X | 3282 hCoV-19/Switzerland/AG-ETHZ-380317/2020\|EPI_ISL_72 | ···················· | ···················· | ···················· | ···················· | ···················· | ···················· |
| X | 3283 hCoV-19/Switzerland/AG-ETHZ-380320/2020\|EPI_ISL_72 | ···················· | ···················· | ···················· | ···················· | ···················· | ···················· |
| X | 3284 hCoV-19/Switzerland/AG-ETHZ-380321/2020\|EPI_ISL_72 | ···················· | ·········A·········· | ···················· | ···················· | ···················· | ···················· |
| X | 3285 hCoV-19/Switzerland/BE-ETHZ-340161/2020\|EPI_ISL_72 | ···················· | ···················· | ···················· | ···················· | ···················· | ···················· |
| X | 3286 hCoV-19/Switzerland/BE-ETHZ-380224/2020\|EPI_ISL_72 | ···················· | ···················· | ···················· | ···················· | ···················· | ···················· |
| X | 3287 hCoV-19/Switzerland/BE-ETHZ-380227/2020\|EPI_ISL_72 | ···················· | ···················· | ···················· | ···················· | ···················· | ···················· |
| X | 3288 hCoV-19/Switzerland/BE-ETHZ-380228/2020\|EPI_ISL_72 | ···················· | ···················· | ···················· | ···················· | ···················· | ···················· |
| X | 3289 hCoV-19/Switzerland/BE-ETHZ-380229/2020\|EPI_ISL_72 | ···················· | ······A············· | ···················· | ···················· | ···················· | ···················· |
| X | 3290 hCoV-19/Switzerland/BE-ETHZ-380280/2020\|EPI_ISL_72 | ···················· | ···················· | ···················· | ···················· | ···················· | ···················· |
| X | 3291 hCoV-19/Switzerland/BE-ETHZ-380284/2020\|EPI_ISL_72 | ···················· | ···················· | ···················· | ···················· | ···················· | ···················· |
| X | 3292 hCoV-19/Switzerland/BE-ETHZ-380306/2020\|EPI_ISL_72 | ···················· | ···················· | ···················· | ···················· | ······K············· | ···················· |
| X | 3293 hCoV-19/Switzerland/BE-ETHZ-380307/2020\|EPI_ISL_72 | ···················· | ···················· | ···················· | ···················· | ···················· | ···················· |
| X | 3294 hCoV-19/Switzerland/BE-ETHZ-380315/2020\|EPI_ISL_72 | ···················· | ···················· | ···················· | ···················· | ···················· | ···················· |
| X | 3295 hCoV-19/Switzerland/BE-ETHZ-380316/2020\|EPI_ISL_72 | ···················· | ···················· | ···················· | ···················· | ···················· | ···················· |

| | | | | | | | |
|---|---|---|---|---|---|---|---|
| X | 3296 hCoV-19/Switzerland/BE-ETHZ-380341/2020\|EPI_ISL_72 | ···················· | ···················· | ····················· | ···················· | ···················· | ···················· |
| X | 3297 hCoV-19/Switzerland/BE-ETHZ-380355/2020\|EPI_ISL_72 | ···················· | ···················· | ····················· | ···················· | ···················· | ···················· |
| X | 3298 hCoV-19/Switzerland/BE-ETHZ-380357/2020\|EPI_ISL_72 | ···················· | ···················· | ····················· | ···················· | ···················· | ···················· |
| X | 3299 hCoV-19/Switzerland/BE-ETHZ-380402/2020\|EPI_ISL_72 | ···················· | ···················· | ····················· | ···················· | ···················· | ···················· |
| X | 3300 hCoV-19/Switzerland/BE-ETHZ-390328/2020\|EPI_ISL_72 | ···················· | ·A·················· | ····················· | ···················· | ···················· | ···················· |
| X | 3301 hCoV-19/Switzerland/BE-ETHZ-390391/2020\|EPI_ISL_72 | ···················· | ·A·················· | ····················· | ···················· | ···················· | ···A················ |
| X | 3302 hCoV-19/Switzerland/BL-ETHZ-340086/2020\|EPI_ISL_72 | ···················· | ···················· | ····················· | ···················· | ···················· | ···················· |
| X | 3303 hCoV-19/Switzerland/BL-ETHZ-340140/2020\|EPI_ISL_72 | ···················· | ···················· | ····················· | ···················· | ···················· | ···················· |
| X | 3304 hCoV-19/Switzerland/BL-ETHZ-340145/2020\|EPI_ISL_72 | ···················· | ···················· | ····················· | ···················· | ···················· | ···················· |
| X | 3305 hCoV-19/Switzerland/BL-ETHZ-340147/2020\|EPI_ISL_72 | ···················· | ···················· | ····················· | ···················· | ···················· | ···················· |
| X | 3306 hCoV-19/Switzerland/BL-ETHZ-340150/2020\|EPI_ISL_72 | ···················· | ···················· | ····················· | ···················· | ···················· | ···················· |
| X | 3307 hCoV-19/Switzerland/BL-ETHZ-380267/2020\|EPI_ISL_72 | ···················· | ···················· | ····················· | ···················· | ···················· | ···················· |
| X | 3308 hCoV-19/Switzerland/BL-ETHZ-380268/2020\|EPI_ISL_72 | ···················· | ···················· | ····················· | ···················· | ···················· | ···················· |
| X | 3309 hCoV-19/Switzerland/BL-ETHZ-380269/2020\|EPI_ISL_72 | ···················· | ···················· | ····················· | ···················· | ···················· | ···················· |
| X | 3310 hCoV-19/Switzerland/BL-ETHZ-380272/2020\|EPI_ISL_72 | ···················· | ···················· | ····················· | ···················· | ···················· | ···················· |
| X | 3311 hCoV-19/Switzerland/BL-ETHZ-380273/2020\|EPI_ISL_72 | ···················· | ···················· | ····················· | ···················· | ···················· | ···················· |
| X | 3312 hCoV-19/Switzerland/BL-ETHZ-380275/2020\|EPI_ISL_72 | ···················· | ·A·················· | ····················· | ···················· | ···················· | ···A················ |
| X | 3313 hCoV-19/Switzerland/BL-ETHZ-380276/2020\|EPI_ISL_72 | ···················· | ···················· | ····················· | ···················· | ···················· | ···················· |
| X | 3314 hCoV-19/Switzerland/BL-ETHZ-380277/2020\|EPI_ISL_72 | ···················· | ·A·················· | ····················· | ···················· | ···················· | ···A················ |
| X | 3315 hCoV-19/Switzerland/BL-ETHZ-380278/2020\|EPI_ISL_72 | ···················· | ···················· | ····················· | ···················· | ···················· | ···················· |
| X | 3316 hCoV-19/Switzerland/BL-ETHZ-380283/2020\|EPI_ISL_72 | ···················· | ···················· | ····················· | ···················· | ···················· | ···················· |
| X | 3317 hCoV-19/Switzerland/BL-ETHZ-380342/2020\|EPI_ISL_72 | ···················· | ···················· | ····················· | ···················· | ···················· | ···················· |
| X | 3318 hCoV-19/Switzerland/BL-ETHZ-380343/2020\|EPI_ISL_72 | ···················· | ···················· | ····················· | ···················· | ···················· | ···················· |
| X | 3319 hCoV-19/Switzerland/BL-ETHZ-380344/2020\|EPI_ISL_72 | ···················· | ·A·················· | ····················· | ···················· | ···················· | ···A················ |
| X | 3320 hCoV-19/Switzerland/BL-ETHZ-380348/2020\|EPI_ISL_72 | ···················· | ···················· | ····················· | ···················· | ······K············· | ···················· |
| X | 3321 hCoV-19/Switzerland/BL-ETHZ-380349/2020\|EPI_ISL_72 | ···················· | ···················· | ····················· | ···················· | ······K············· | ···················· |
| X | 3322 hCoV-19/Switzerland/BL-ETHZ-380352/2020\|EPI_ISL_72 | ···················· | ···················· | ····················· | ···················· | ···················· | ···················· |
| X | 3323 hCoV-19/Switzerland/BL-ETHZ-380353/2020\|EPI_ISL_72 | ···················· | ···················· | ····················· | ···················· | ···················· | ···················· |
| X | 3324 hCoV-19/Switzerland/BL-ETHZ-380354/2020\|EPI_ISL_72 | ···················· | ·A·················· | ····················· | ···················· | ···················· | ···A················ |
| X | 3325 hCoV-19/Switzerland/BL-ETHZ-380376/2020\|EPI_ISL_72 | ···················· | ···················· | ····················· | ···················· | ···················· | ···················· |
| X | 3326 hCoV-19/Switzerland/BL-ETHZ-380387/2020\|EPI_ISL_72 | ···················· | ···················· | ····················· | ···················· | ···················· | ···················· |
| X | 3327 hCoV-19/Switzerland/BL-ETHZ-380389/2020\|EPI_ISL_72 | ···················· | ···················· | ····················· | ···················· | ···················· | ···················· |
| X | 3328 hCoV-19/Switzerland/BL-ETHZ-380394/2020\|EPI_ISL_72 | ···················· | ·A·················· | ····················· | ···················· | ···················· | ···A················ |
| X | 3329 hCoV-19/Denmark/DCGC-14810/2020\|EPI_ISL_712000\|202 | ···················· | ···················· | ····················· | ···················· | ···················· | ···················· |
| X | 3330 hCoV-19/Denmark/DCGC-14812/2020\|EPI_ISL_712001\|202 | ···················· | ·A·················· | ····················· | ···················· | ···················· | ···A················ |
| X | 3331 hCoV-19/Denmark/DCGC-14815/2020\|EPI_ISL_712003\|202 | ···················· | ···················· | ····················· | ···················· | ···················· | ···················· |
| X | 3332 hCoV-19/Denmark/DCGC-14823/2020\|EPI_ISL_712010\|202 | ···················· | ···················· | ····················· | ···················· | ···················· | ···················· |
| X | 3333 hCoV-19/Denmark/DCGC-14825/2020\|EPI_ISL_712011\|202 | ···················· | ···················· | ····················· | ···················· | ···················· | ···················· |
| X | 3334 hCoV-19/Denmark/DCGC-14828/2020\|EPI_ISL_712012\|202 | ···················· | ···················· | ····················· | ···················· | ···················· | ···················· |
| X | 3335 hCoV-19/Denmark/DCGC-14830/2020\|EPI_ISL_712013\|202 | ···················· | ···················· | ····················· | ···················· | ···················· | ···················· |
| X | 3336 hCoV-19/Denmark/DCGC-14831/2020\|EPI_ISL_712014\|202 | ···················· | ···················· | ····················· | ···················· | ···················· | ···················· |
| X | 3337 hCoV-19/Denmark/DCGC-14832/2020\|EPI_ISL_712015\|202 | ···················· | ···················· | ····················· | ···················· | ···················· | ···················· |
| X | 3338 hCoV-19/Denmark/DCGC-14835/2020\|EPI_ISL_712016\|202 | ···················· | ···················· | ····················· | ···················· | ···················· | ···················· |
| X | 3339 hCoV-19/Denmark/DCGC-14836/2020\|EPI_ISL_712017\|202 | ···················· | ···················· | ····················· | ···················· | ···················· | ···················· |
| X | 3340 hCoV-19/Denmark/DCGC-14837/2020\|EPI_ISL_712018\|202 | ···················· | ···················· | ····················· | ···················· | ···················· | ···················· |
| X | 3341 hCoV-19/Denmark/DCGC-14840/2020\|EPI_ISL_712019\|202 | ···················· | ···················· | ····················· | ···················· | ···················· | ···················· |
| X | 3342 hCoV-19/Denmark/DCGC-14841/2020\|EPI_ISL_712020\|202 | ···················· | ···················· | ····················· | ···················· | ···················· | ···················· |
| X | 3343 hCoV-19/Denmark/DCGC-14843/2020\|EPI_ISL_712021\|202 | ···················· | ···················· | ····················· | ···················· | ···················· | ···················· |
| X | 3344 hCoV-19/Denmark/DCGC-14844/2020\|EPI_ISL_712022\|202 | ···················· | ···················· | ····················· | ···················· | ···················· | ···················· |
| X | 3345 hCoV-19/Denmark/DCGC-14845/2020\|EPI_ISL_712023\|202 | ···················· | ···················· | ····················· | ···················· | ···················· | ···················· |
| X | 3346 hCoV-19/Denmark/DCGC-14848/2020\|EPI_ISL_712024\|202 | ···················· | ···················· | ····················· | ···················· | ···················· | ···················· |
| X | 3347 hCoV-19/Denmark/DCGC-14849/2020\|EPI_ISL_712025\|202 | ···················· | ···················· | ····················· | ···················· | ···················· | ···················· |
| X | 3348 hCoV-19/Denmark/DCGC-14850/2020\|EPI_ISL_712026\|202 | ···················· | ···················· | ····················· | ···················· | ······T············· | ···················· |
| X | 3349 hCoV-19/Denmark/DCGC-14852/2020\|EPI_ISL_712027\|202 | ···················· | ···················· | ····················· | ···················· | ···················· | ···················· |
| X | 3350 hCoV-19/Denmark/DCGC-14853/2020\|EPI_ISL_712028\|202 | ···················· | ···················· | ····················· | ···················· | ···················· | ···················· |
| X | 3351 hCoV-19/Denmark/DCGC-14854/2020\|EPI_ISL_712029\|202 | ···················· | ·A·················· | ····················· | ···················· | ···················· | ···················· |
| X | 3352 hCoV-19/Denmark/DCGC-14855/2020\|EPI_ISL_712030\|202 | ···················· | ···················· | ····················· | ···················· | ···················· | ···················· |
| X | 3353 hCoV-19/Denmark/DCGC-14856/2020\|EPI_ISL_712031\|202 | ···················· | ···················· | ····················· | ···················· | ···················· | ···················· |
| X | 3354 hCoV-19/Denmark/DCGC-14858/2020\|EPI_ISL_712032\|202 | ···················· | ···················· | ····················· | ···················· | ···················· | ···················· |
| X | 3355 hCoV-19/Denmark/DCGC-14860/2020\|EPI_ISL_712033\|202 | ···················· | ···················· | ····················· | ···················· | ···················· | ···················· |
| X | 3356 hCoV-19/Denmark/DCGC-14861/2020\|EPI_ISL_712034\|202 | ···················· | ···················· | ····················· | ···················· | ···················· | ···················· |
| X | 3357 hCoV-19/Denmark/DCGC-14863/2020\|EPI_ISL_712035\|202 | ···················· | ···················· | ····················· | ···················· | ···················· | ···················· |
| X | 3358 hCoV-19/Denmark/DCGC-14864/2020\|EPI_ISL_712036\|202 | ···················· | ···················· | ····················· | ···················· | ···················· | ···················· |
| X | 3359 hCoV-19/Denmark/DCGC-14865/2020\|EPI_ISL_712037\|202 | ···················· | ···················· | ····················· | ···················· | ···················· | ···················· |
| X | 3360 hCoV-19/Denmark/DCGC-14866/2020\|EPI_ISL_712038\|202 | ···················· | ···················· | ····················· | ···················· | ···················· | ···················· |
| X | 3361 hCoV-19/Denmark/DCGC-14867/2020\|EPI_ISL_712039\|202 | ···················· | ···················· | ····················· | ···················· | ···················· | ···················· |
| X | 3362 hCoV-19/Denmark/DCGC-14868/2020\|EPI_ISL_712040\|202 | ···················· | ···················· | ····················· | ···················· | ···················· | ···················· |
| X | 3363 hCoV-19/Denmark/DCGC-14874/2020\|EPI_ISL_712041\|202 | ···················· | ···················· | ····················· | ···················· | ···················· | ···················· |
| X | 3364 hCoV-19/Denmark/DCGC-14875/2020\|EPI_ISL_712042\|202 | ···················· | ···················· | ····················· | ···················· | ···················· | ···················· |
| X | 3365 hCoV-19/Denmark/DCGC-14880/2020\|EPI_ISL_712043\|202 | ···················· | ···················· | ····················· | ···················· | ···················· | ···················· |
| X | 3366 hCoV-19/Denmark/DCGC-14882/2020\|EPI_ISL_712044\|202 | ···················· | ···················· | ····················· | ···················· | ···················· | ···················· |
| X | 3367 hCoV-19/Denmark/DCGC-14885/2020\|EPI_ISL_712045\|202 | ···················· | ···················· | ····················· | ···················· | ···················· | ···················· |
| X | 3368 hCoV-19/Denmark/DCGC-14887/2020\|EPI_ISL_712046\|202 | ···················· | ···················· | ····················· | ···················· | ···················· | ···················· |
| X | 3369 hCoV-19/Denmark/DCGC-14888/2020\|EPI_ISL_712047\|202 | ···················· | ···················· | ····················· | ···················· | ···················· | ···················· |
| X | 3370 hCoV-19/Denmark/DCGC-14889/2020\|EPI_ISL_712048\|202 | ···················· | ···················· | ····················· | ···················· | ···················· | ···················· |
| X | 3371 hCoV-19/Denmark/DCGC-14890/2020\|EPI_ISL_712049\|202 | ···················· | ···················· | ····················· | ···················· | ···················· | ···················· |
| X | 3372 hCoV-19/Denmark/DCGC-14891/2020\|EPI_ISL_712050\|202 | ···················· | ···················· | ····················· | ···················· | ···················· | ···················· |
| X | 3373 hCoV-19/Denmark/DCGC-14892/2020\|EPI_ISL_712051\|202 | ···················· | ···················· | ····················· | ···················· | ···················· | ···················· |
| X | 3374 hCoV-19/Denmark/DCGC-14895/2020\|EPI_ISL_712052\|202 | ···················· | ···················· | ····················· | ···················· | ···················· | ···················· |
| X | 3375 hCoV-19/Denmark/DCGC-14896/2020\|EPI_ISL_712053\|202 | ···················· | ···················· | ····················· | ···················· | ···················· | ···················· |

| | | | | | | | |
|---|---|---|---|---|---|---|---|
| X | 3376 hCoV-19/Denmark/DCGC-14897/2020\|EPI_ISL_712054\|202 | ···················· | ···················· | ···················· | ···················· | ···················· | ···················· |
| X | 3377 hCoV-19/Denmark/DCGC-14902/2020\|EPI_ISL_712055\|202 | ···················· | ···················· | ···················· | ···················· | ···················· | ···················· |
| X | 3378 hCoV-19/Denmark/DCGC-14904/2020\|EPI_ISL_712056\|202 | ···················· | ···················· | ···················· | ···················· | ···················· | ···················· |
| X | 3379 hCoV-19/Denmark/DCGC-14905/2020\|EPI_ISL_712057\|202 | ···················· | ···················· | ···················· | ···················· | ···················· | ···················· |
| X | 3380 hCoV-19/Denmark/DCGC-14906/2020\|EPI_ISL_712058\|202 | ···················· | ···················· | ···················· | ···················· | ···················· | ···················· |
| X | 3381 hCoV-19/Denmark/DCGC-14908/2020\|EPI_ISL_712059\|202 | ···················· | ···················· | ···················· | ···················· | ···················· | ···················· |
| X | 3382 hCoV-19/Tunisia/ADAGE-18505/2020\|EPI_ISL_712060\|20 | ···················· | ···················· | ···················· | ···················· | ···················· | ···················· |
| X | 3383 hCoV-19/Tunisia/ADAGE-24755/2020\|EPI_ISL_712062\|20 | ···················· | ···················· | ···················· | ···················· | ···················· | ···················· |
| X | 3384 hCoV-19/Tunisia/ADAGE-24758/2020\|EPI_ISL_712063\|20 | ···················· | ······A············· | ···················· | ···················· | ···················· | ···················· |
| X | 3385 hCoV-19/Tunisia/ADAGE-24763/2020\|EPI_ISL_712064\|20 | ···················· | ···················· | ···················· | ···················· | ···················· | ···················· |
| X | 3386 hCoV-19/Tunisia/ADAGE-24776/2020\|EPI_ISL_712065\|20 | ···················· | ···················· | ···················· | ···················· | ···················· | ···················· |
| X | 3387 hCoV-19/Tunisia/ADAGE-24779/2020\|EPI_ISL_712066\|20 | ···················· | ···················· | ···················· | ···················· | ···················· | ···················· |
| X | 3388 hCoV-19/Tunisia/ADAGE-24804/2020\|EPI_ISL_712067\|20 | ···················· | ···················· | ···················· | ···················· | ···················· | ···················· |
| X | 3389 hCoV-19/Denmark/DCGC-14970/2020\|EPI_ISL_713315\|202 | ···················· | ···················· | ···················· | ···················· | ···················· | ···················· |
| X | 3390 hCoV-19/Denmark/DCGC-14979/2020\|EPI_ISL_713316\|202 | ···················· | ···················· | ···················· | ···················· | ···················· | ···················· |
| X | 3391 hCoV-19/Denmark/DCGC-15072/2020\|EPI_ISL_713317\|202 | ···················· | ···················· | ···················· | ···················· | ···················· | ···················· |
| X | 3392 hCoV-19/Denmark/DCGC-15093/2020\|EPI_ISL_713318\|202 | ···················· | ····A··············· | ···················· | ···················· | ···················· | ···················· |
| X | 3393 hCoV-19/Denmark/DCGC-14913/2020\|EPI_ISL_713331\|202 | ···················· | ···················· | ···················· | ···················· | ···················· | ···················· |
| X | 3394 hCoV-19/Denmark/DCGC-14925/2020\|EPI_ISL_713332\|202 | ···················· | ···················· | ···················· | ···················· | ···················· | ···················· |
| X | 3395 hCoV-19/Denmark/DCGC-14928/2020\|EPI_ISL_713333\|202 | ···················· | ···················· | ···················· | ···················· | ···················· | ···················· |
| X | 3396 hCoV-19/Denmark/DCGC-15034/2020\|EPI_ISL_713340\|202 | ···················· | ···················· | ···················· | ···················· | ···················· | ···················· |
| X | 3397 hCoV-19/Denmark/DCGC-15040/2020\|EPI_ISL_713341\|202 | ···················· | ···················· | ···················· | ···················· | ···················· | ···················· |
| X | 3398 hCoV-19/Denmark/DCGC-15049/2020\|EPI_ISL_713342\|202 | ···················· | ···················· | ···················· | ···················· | ···················· | ···················· |
| X | 3399 hCoV-19/Denmark/DCGC-15053/2020\|EPI_ISL_713343\|202 | ···················· | ···················· | ···················· | ···················· | ···················· | ···················· |
| X | 3400 hCoV-19/Denmark/DCGC-15055/2020\|EPI_ISL_713344\|202 | ···················· | ···················· | ···················· | ···················· | ···················· | ···················· |
| X | 3401 hCoV-19/Denmark/DCGC-15062/2020\|EPI_ISL_713345\|202 | ···················· | ···················· | ···················· | ···················· | ···················· | ···················· |
| X | 3402 hCoV-19/Denmark/DCGC-15085/2020\|EPI_ISL_713346\|202 | ···················· | ···················· | ···················· | ···················· | ···················· | ···················· |
| X | 3403 hCoV-19/Denmark/DCGC-15086/2020\|EPI_ISL_713347\|202 | ···················· | ···················· | ···················· | ···················· | ···················· | ···················· |
| X | 3404 hCoV-19/Denmark/DCGC-15087/2020\|EPI_ISL_713348\|202 | ···················· | ···················· | ···················· | ···················· | ···················· | ···················· |
| X | 3405 hCoV-19/Denmark/DCGC-15091/2020\|EPI_ISL_713349\|202 | ···················· | ···················· | ···················· | ···················· | ···················· | ···················· |
| X | 3406 hCoV-19/Denmark/DCGC-15092/2020\|EPI_ISL_713350\|202 | ···················· | ···················· | ···················· | ···················· | ···················· | ···················· |
| X | 3407 hCoV-19/Denmark/DCGC-15105/2020\|EPI_ISL_713351\|202 | ···················· | ···················· | ···················· | ···················· | ···················· | ···················· |
| X | 3408 hCoV-19/Denmark/DCGC-15110/2020\|EPI_ISL_713352\|202 | ···················· | ···················· | ···················· | ···················· | ···················· | ···················· |
| X | 3409 hCoV-19/Denmark/DCGC-15116/2020\|EPI_ISL_713353\|202 | ···················· | ···················· | ···················· | ···················· | ···················· | ···················· |
| X | 3410 hCoV-19/Denmark/DCGC-15117/2020\|EPI_ISL_713354\|202 | ···················· | ···················· | ···················· | ···················· | ···················· | ···················· |
| X | 3411 hCoV-19/Denmark/DCGC-15120/2020\|EPI_ISL_713355\|202 | ···················· | ···················· | ···················· | ···················· | ···················· | ···················· |
| X | 3412 hCoV-19/Denmark/DCGC-15123/2020\|EPI_ISL_713356\|202 | ···················· | ···················· | ···················· | ···················· | ···················· | ···················· |
| X | 3413 hCoV-19/Denmark/DCGC-15188/2020\|EPI_ISL_713357\|202 | ···················· | ···················· | ···················· | ···················· | ···················· | ···················· |
| X | 3414 hCoV-19/Denmark/DCGC-15192/2020\|EPI_ISL_713358\|202 | ···················· | ···················· | ···················· | ···················· | ···················· | ···················· |
| X | 3415 hCoV-19/Denmark/DCGC-15204/2020\|EPI_ISL_713359\|202 | ···················· | ···················· | ···················· | ···················· | ···················· | ···················· |
| X | 3416 hCoV-19/Denmark/DCGC-15206/2020\|EPI_ISL_713360\|202 | ···················· | ···················· | ···················· | ···················· | ···················· | ···················· |
| X | 3417 hCoV-19/Denmark/DCGC-15209/2020\|EPI_ISL_713361\|202 | ···················· | ···················· | ···················· | ···················· | ···················· | ···················· |
| X | 3418 hCoV-19/Denmark/DCGC-15229/2020\|EPI_ISL_713362\|202 | ···················· | ···················· | ···················· | ···················· | ···················· | ···················· |
| X | 3419 hCoV-19/Denmark/DCGC-15243/2020\|EPI_ISL_713363\|202 | ···················· | ···················· | ···················· | ···················· | ···················· | ···················· |
| X | 3420 hCoV-19/Denmark/DCGC-15244/2020\|EPI_ISL_713364\|202 | ···················· | ···················· | ···················· | ···················· | ···················· | ···················· |
| X | 3421 hCoV-19/Denmark/DCGC-15278/2020\|EPI_ISL_713365\|202 | ···················· | ···················· | ···················· | ···················· | ···················· | ···················· |
| X | 3422 hCoV-19/Denmark/DCGC-15287/2020\|EPI_ISL_713366\|202 | ···················· | ···················· | ···················· | ···················· | ···················· | ···················· |
| X | 3423 hCoV-19/Denmark/DCGC-15293/2020\|EPI_ISL_713367\|202 | ···················· | ···················· | ···················· | ···················· | ···················· | ···················· |
| X | 3424 hCoV-19/Denmark/DCGC-15303/2020\|EPI_ISL_713368\|202 | ···················· | ···················· | ···················· | ···················· | ···················· | ···················· |
| X | 3425 hCoV-19/Denmark/DCGC-15309/2020\|EPI_ISL_713369\|202 | ···················· | ···················· | ···················· | ···················· | ···················· | ···················· |
| X | 3426 hCoV-19/Denmark/DCGC-15327/2020\|EPI_ISL_713370\|202 | ···················· | ···················· | ···················· | ···················· | ···················· | ···················· |
| X | 3427 hCoV-19/Denmark/DCGC-15335/2020\|EPI_ISL_713371\|202 | ···················· | ···················· | ···················· | ···················· | ···················· | ···················· |
| X | 3428 hCoV-19/Denmark/DCGC-15336/2020\|EPI_ISL_713372\|202 | ···················· | ···················· | ···················· | ···················· | ···················· | ···················· |
| X | 3429 hCoV-19/Denmark/DCGC-15346/2020\|EPI_ISL_713373\|202 | ···················· | ···················· | ···················· | ···················· | ···················· | ···················· |
| X | 3430 hCoV-19/Denmark/DCGC-15355/2020\|EPI_ISL_713374\|202 | ···················· | ···················· | ···················· | ···················· | ···················· | ···················· |
| X | 3431 hCoV-19/Denmark/DCGC-15360/2020\|EPI_ISL_713375\|202 | ···················· | ···················· | ···················· | ···················· | ···················· | ···················· |
| X | 3432 hCoV-19/Denmark/DCGC-15372/2020\|EPI_ISL_713376\|202 | ···················· | ···················· | ···················· | ···················· | ···················· | ···················· |
| X | 3433 hCoV-19/Denmark/DCGC-15382/2020\|EPI_ISL_713377\|202 | ···················· | ···················· | ···················· | ···················· | ···················· | ···················· |
| X | 3434 hCoV-19/Denmark/DCGC-15401/2020\|EPI_ISL_713378\|202 | ···················· | ···················· | ···················· | ···················· | ···················· | ···················· |
| X | 3435 hCoV-19/Denmark/DCGC-15407/2020\|EPI_ISL_713379\|202 | ···················· | ···················· | ···················· | ···················· | ···················· | ···················· |
| X | 3436 hCoV-19/Denmark/DCGC-15436/2020\|EPI_ISL_713380\|202 | ···················· | ···················· | ···················· | ···················· | ···················· | ···················· |
| X | 3437 hCoV-19/Denmark/DCGC-15442/2020\|EPI_ISL_713381\|202 | ···················· | ···················· | ···················· | ···················· | ···················· | ···A················ |
| X | 3438 hCoV-19/Denmark/DCGC-15445/2020\|EPI_ISL_713382\|202 | ···················· | ···················· | ···················· | ···················· | ···················· | ···················· |
| X | 3439 hCoV-19/Denmark/DCGC-15448/2020\|EPI_ISL_713383\|202 | ···················· | ···················· | ···················· | ···················· | ···················· | ···················· |
| X | 3440 hCoV-19/Denmark/DCGC-15465/2020\|EPI_ISL_713384\|202 | ···················· | ···················· | ···················· | ···················· | ···················· | ···················· |
| X | 3441 hCoV-19/Denmark/DCGC-15468/2020\|EPI_ISL_713385\|202 | ···················· | ···················· | ···················· | ···················· | ···················· | ···················· |
| X | 3442 hCoV-19/Denmark/DCGC-15472/2020\|EPI_ISL_713386\|202 | ···················· | ···················· | ···················· | ···················· | ···················· | ···················· |
| X | 3443 hCoV-19/Denmark/DCGC-15488/2020\|EPI_ISL_713387\|202 | ···················· | ···················· | ···················· | ···················· | ···················· | ···················· |
| X | 3444 hCoV-19/Denmark/DCGC-15493/2020\|EPI_ISL_713388\|202 | ···················· | ···················· | ···················· | ···················· | ···················· | ···················· |
| X | 3445 hCoV-19/Denmark/DCGC-15510/2020\|EPI_ISL_713389\|202 | ···················· | ···················· | ···················· | ···················· | ···················· | ···················· |
| X | 3446 hCoV-19/Denmark/DCGC-15530/2020\|EPI_ISL_713390\|202 | ···················· | ···················· | ···················· | ···················· | ···················· | ···················· |
| X | 3447 hCoV-19/Denmark/DCGC-15540/2020\|EPI_ISL_713391\|202 | ···················· | ···················· | ···················· | ···················· | ···················· | ···················· |
| X | 3448 hCoV-19/Denmark/DCGC-15541/2020\|EPI_ISL_713392\|202 | ···················· | ···················· | ···················· | ···················· | ···················· | ···················· |
| X | 3449 hCoV-19/Denmark/DCGC-15553/2020\|EPI_ISL_713393\|202 | ···················· | ···················· | ···················· | ···················· | ···················· | ···················· |
| X | 3450 hCoV-19/Denmark/DCGC-15567/2020\|EPI_ISL_713394\|202 | ···················· | ···················· | ···················· | ···················· | ···················· | ···················· |
| X | 3451 hCoV-19/Denmark/DCGC-15572/2020\|EPI_ISL_713395\|202 | ···················· | ···················· | ···················· | ···················· | ···················· | ···················· |
| X | 3452 hCoV-19/Denmark/DCGC-15580/2020\|EPI_ISL_713396\|202 | ···················· | ···················· | ···················· | ···················· | ···················· | ···················· |
| X | 3453 hCoV-19/Denmark/DCGC-15588/2020\|EPI_ISL_713397\|202 | ···················· | ···················· | ···················· | ···················· | ···················· | ···················· |
| X | 3454 hCoV-19/Denmark/DCGC-15589/2020\|EPI_ISL_713398\|202 | ···················· | ···················· | ···················· | ···················· | ···················· | ···················· |
| X | 3455 hCoV-19/Denmark/DCGC-15603/2020\|EPI_ISL_713399\|202 | ···················· | ···················· | ···················· | ···················· | ···················· | ···················· |

| | | | | | | | |
|---|---|---|---|---|---|---|---|
| X | 3456 hCoV-19/Denmark/DCGC-16189/2020\|EPI_ISL_714628\|202 | ···················· | ···················· | ···················· | ···················· | ···················· | ···················· |
| X | 3457 hCoV-19/Denmark/DCGC-16190/2020\|EPI_ISL_714629\|202 | ···················· | ···················· | ···················· | ···················· | ···················· | ···················· |
| X | 3458 hCoV-19/Denmark/DCGC-16191/2020\|EPI_ISL_714630\|202 | ···················· | ···················· | ···················· | ···················· | ···················· | ···················· |
| X | 3459 hCoV-19/Denmark/DCGC-16193/2020\|EPI_ISL_714631\|202 | ···················· | ···················· | ···················· | ···················· | ···················· | ···················· |
| X | 3460 hCoV-19/Denmark/DCGC-16196/2020\|EPI_ISL_714632\|202 | ···················· | ···················· | ···················· | ···················· | ···················· | ···················· |
| X | 3461 hCoV-19/Denmark/DCGC-16201/2020\|EPI_ISL_714633\|202 | ···················· | ···················· | ···················· | ···················· | ···················· | ···················· |
| X | 3462 hCoV-19/Denmark/DCGC-16202/2020\|EPI_ISL_714634\|202 | ···················· | ···················· | ···················· | ···················· | ···················· | ···················· |
| X | 3463 hCoV-19/Denmark/DCGC-16203/2020\|EPI_ISL_714635\|202 | ···················· | ···················· | ···················· | ···················· | ···················· | ···················· |
| X | 3464 hCoV-19/Denmark/DCGC-16205/2020\|EPI_ISL_714636\|202 | ···················· | ···················· | ···················· | ···················· | ···················· | ···················· |
| X | 3465 hCoV-19/Denmark/DCGC-16206/2020\|EPI_ISL_714637\|202 | ···················· | ···················· | ···················· | ···················· | ···················· | ···················· |
| X | 3466 hCoV-19/Denmark/DCGC-16207/2020\|EPI_ISL_714638\|202 | ···················· | ···················· | ···················· | ···················· | ···················· | ···················· |
| X | 3467 hCoV-19/Denmark/DCGC-16208/2020\|EPI_ISL_714639\|202 | ···················· | ···················· | ···················· | ···················· | ···················· | ···················· |
| X | 3468 hCoV-19/Denmark/DCGC-16209/2020\|EPI_ISL_714640\|202 | ···················· | ···················· | ···················· | ···················· | ···················· | ···················· |
| X | 3469 hCoV-19/Denmark/DCGC-16211/2020\|EPI_ISL_714641\|202 | ···················· | ···················· | ···················· | ···················· | ···················· | ···················· |
| X | 3470 hCoV-19/Denmark/DCGC-16213/2020\|EPI_ISL_714642\|202 | ···················· | ···A················ | ···················· | ···················· | ···················· | ···················· |
| X | 3471 hCoV-19/Denmark/DCGC-16214/2020\|EPI_ISL_714643\|202 | ···················· | ···················· | ···················· | ···················· | ···················· | ···················· |
| X | 3472 hCoV-19/Denmark/DCGC-16218/2020\|EPI_ISL_714644\|202 | ···················· | ···················· | ···················· | ···················· | ···················· | ···················· |
| X | 3473 hCoV-19/Denmark/DCGC-16219/2020\|EPI_ISL_714645\|202 | ···················· | ···················· | ···················· | ···················· | ···················· | ···················· |
| X | 3474 hCoV-19/Denmark/DCGC-16220/2020\|EPI_ISL_714646\|202 | ···················· | ···················· | ···················· | ···················· | ···················· | ···················· |
| X | 3475 hCoV-19/Denmark/DCGC-16221/2020\|EPI_ISL_714647\|202 | ···················· | ···················· | ···················· | ···················· | ···················· | ···················· |
| X | 3476 hCoV-19/Denmark/DCGC-16224/2020\|EPI_ISL_714648\|202 | ···················· | ···················· | ···················· | ···················· | ···················· | ···················· |
| X | 3477 hCoV-19/Denmark/DCGC-16225/2020\|EPI_ISL_714649\|202 | ···················· | ·A·················· | ···················· | ···················· | ···················· | ···················· |
| X | 3478 hCoV-19/Denmark/DCGC-16240/2020\|EPI_ISL_714660\|202 | ···················· | ·A·················· | ···················· | ···················· | ···················· | ···················· |
| X | 3479 hCoV-19/Denmark/DCGC-16243/2020\|EPI_ISL_714661\|202 | ···················· | ···················· | ···················· | ···················· | ···················· | ···················· |
| X | 3480 hCoV-19/Denmark/DCGC-16246/2020\|EPI_ISL_714662\|202 | ···················· | ···················· | ···················· | ···················· | ···················· | ···················· |
| X | 3481 hCoV-19/Denmark/DCGC-16247/2020\|EPI_ISL_714663\|202 | ···················· | ···················· | ···················· | ···················· | ·····T·············· | ···················· |
| X | 3482 hCoV-19/Denmark/DCGC-16259/2020\|EPI_ISL_714670\|202 | ···················· | ···················· | ···················· | ···················· | ···················· | ···················· |
| X | 3483 hCoV-19/Denmark/DCGC-16263/2020\|EPI_ISL_714671\|202 | ···················· | ···················· | ···················· | ···················· | ···················· | ···················· |
| X | 3484 hCoV-19/Denmark/DCGC-16266/2020\|EPI_ISL_714672\|202 | ···················· | ···················· | ···················· | ···················· | ···················· | ···················· |
| X | 3485 hCoV-19/Denmark/DCGC-16267/2020\|EPI_ISL_714673\|202 | ···················· | ···················· | ···················· | ···················· | ···················· | ···················· |
| X | 3486 hCoV-19/Denmark/DCGC-16270/2020\|EPI_ISL_714674\|202 | ···················· | ···················· | ···················· | ···················· | ···················· | ···················· |
| X | 3487 hCoV-19/Denmark/DCGC-16271/2020\|EPI_ISL_714675\|202 | ···················· | ···················· | ···················· | ···················· | ···················· | ···················· |
| X | 3488 hCoV-19/Denmark/DCGC-16273/2020\|EPI_ISL_714676\|202 | ···················· | ···················· | ···················· | ···················· | ···················· | ···················· |
| X | 3489 hCoV-19/Denmark/DCGC-16274/2020\|EPI_ISL_714677\|202 | ···················· | ···················· | ···················· | ···················· | ···················· | ···················· |
| X | 3490 hCoV-19/Denmark/DCGC-16276/2020\|EPI_ISL_714678\|202 | ···················· | ···················· | ···················· | ···················· | ·····T·············· | ···················· |
| X | 3491 hCoV-19/Denmark/DCGC-16278/2020\|EPI_ISL_714679\|202 | ···················· | ···················· | ···················· | ···················· | ···················· | ···················· |
| X | 3492 hCoV-19/Denmark/DCGC-16281/2020\|EPI_ISL_714680\|202 | ···················· | ···················· | ···················· | ···················· | ···················· | ···················· |
| X | 3493 hCoV-19/Denmark/DCGC-16282/2020\|EPI_ISL_714681\|202 | ···················· | ···················· | ···················· | ···················· | ···················· | ···················· |
| X | 3494 hCoV-19/Denmark/DCGC-16286/2020\|EPI_ISL_714682\|202 | ···················· | ···················· | ···················· | ···················· | ···················· | ···················· |
| X | 3495 hCoV-19/Denmark/DCGC-16288/2020\|EPI_ISL_714683\|202 | ···················· | ···················· | ···················· | ···················· | ·····T·············· | ···················· |
| X | 3496 hCoV-19/Denmark/DCGC-16289/2020\|EPI_ISL_714684\|202 | ···················· | ·A·················· | ···················· | ···················· | ···················· | ··A················· |
| X | 3497 hCoV-19/Denmark/DCGC-16291/2020\|EPI_ISL_714685\|202 | ···················· | ···················· | ···················· | ···················· | ···················· | ···················· |
| X | 3498 hCoV-19/Denmark/DCGC-16297/2020\|EPI_ISL_714686\|202 | ···················· | ···················· | ···················· | ···················· | ···················· | ···················· |
| X | 3499 hCoV-19/Denmark/DCGC-16298/2020\|EPI_ISL_714687\|202 | ···················· | ···················· | ···················· | ···················· | ···················· | ···················· |
| X | 3500 hCoV-19/Denmark/DCGC-16299/2020\|EPI_ISL_714688\|202 | ···················· | ···················· | ···················· | ···················· | ···················· | ···················· |
| X | 3501 hCoV-19/Denmark/DCGC-16300/2020\|EPI_ISL_714689\|202 | ···················· | ···················· | ···················· | ···················· | ···················· | ···················· |
| X | 3502 hCoV-19/Denmark/DCGC-16301/2020\|EPI_ISL_714690\|202 | ···················· | ···················· | ···················· | ···················· | ···················· | ···················· |
| X | 3503 hCoV-19/Denmark/DCGC-16302/2020\|EPI_ISL_714691\|202 | ···················· | ···················· | ···················· | ···················· | ···················· | ···················· |
| X | 3504 hCoV-19/Denmark/DCGC-16303/2020\|EPI_ISL_714692\|202 | ···················· | ···················· | ···················· | ···················· | ···················· | ···················· |
| X | 3505 hCoV-19/Denmark/DCGC-16304/2020\|EPI_ISL_714693\|202 | ···················· | ···················· | ···················· | ···················· | ···················· | ···················· |
| X | 3506 hCoV-19/Denmark/DCGC-16306/2020\|EPI_ISL_714694\|202 | ···················· | ···················· | ···················· | ···················· | ···················· | ···················· |
| X | 3507 hCoV-19/Denmark/DCGC-16308/2020\|EPI_ISL_714695\|202 | ···················· | ···················· | ···················· | ···················· | ···················· | ···················· |
| X | 3508 hCoV-19/Denmark/DCGC-16310/2020\|EPI_ISL_714696\|202 | ···················· | ···················· | ···················· | ···················· | ···················· | ···················· |
| X | 3509 hCoV-19/Denmark/DCGC-16313/2020\|EPI_ISL_714697\|202 | ···················· | ···················· | ···················· | ···················· | ···················· | ···················· |
| X | 3510 hCoV-19/Denmark/DCGC-16314/2020\|EPI_ISL_714698\|202 | ···················· | ···················· | ···················· | ···················· | ···················· | ···················· |
| X | 3511 hCoV-19/Denmark/DCGC-16316/2020\|EPI_ISL_714699\|202 | ···················· | ···················· | ···················· | ···················· | ···················· | ···················· |
| X | 3512 hCoV-19/England/CAMC-C3FBED/2020\|EPI_ISL_719500\|20 | ···················· | ···················· | ···················· | ···················· | ···················· | ···················· |
| X | 3513 hCoV-19/England/CAMC-C43E83/2020\|EPI_ISL_719505\|20 | ···················· | ···················· | ···················· | ···················· | ···················· | ···················· |
| X | 3514 hCoV-19/England/CAMC-C440AA/2020\|EPI_ISL_719509\|20 | ···················· | ·A·················· | ···················· | ···················· | ···················· | ··A················· |
| X | 3515 hCoV-19/England/CAMC-C3FC26/2020\|EPI_ISL_719517\|20 | ···················· | ···················· | ···················· | ···················· | ···················· | ···················· |
| X | 3516 hCoV-19/England/MILK-BB1A41/2020\|EPI_ISL_720500\|20 | ···················· | ···················· | ···················· | ···················· | ···················· | ··A················· |
| X | 3517 hCoV-19/England/MILK-BB0E82/2020\|EPI_ISL_720501\|20 | ···················· | ···················· | ···················· | ···················· | ···················· | ···················· |
| X | 3518 hCoV-19/Switzerland/GE-ETHZ-340105/2020\|EPI_ISL_72 | ···················· | ···················· | ···················· | ···················· | ···················· | ···················· |
| X | 3519 hCoV-19/Switzerland/GE-ETHZ-340107/2020\|EPI_ISL_72 | ···················· | ···················· | ···················· | ···················· | ···················· | ···················· |
| X | 3520 hCoV-19/Switzerland/GE-ETHZ-380213/2020\|EPI_ISL_72 | ···················· | ···················· | ···················· | ···················· | ···················· | ···················· |
| X | 3521 hCoV-19/Switzerland/GE-ETHZ-380218/2020\|EPI_ISL_72 | ···················· | ···················· | ···················· | ···················· | ···················· | ···················· |
| X | 3522 hCoV-19/Switzerland/GR-ETHZ-340106/2020\|EPI_ISL_72 | ···················· | ···················· | ···················· | ···················· | ···················· | ···················· |
| X | 3523 hCoV-19/Switzerland/GR-ETHZ-340131/2020\|EPI_ISL_72 | ···················· | ···················· | ···················· | ···················· | ···················· | ···················· |
| X | 3524 hCoV-19/Switzerland/GR-ETHZ-340133/2020\|EPI_ISL_72 | ···················· | ···················· | ···················· | ···················· | ···················· | ···················· |
| X | 3525 hCoV-19/Switzerland/GR-ETHZ-390381/2020\|EPI_ISL_72 | ···················· | ···················· | ···················· | ···················· | ···················· | ···················· |
| X | 3526 hCoV-19/Switzerland/GR-ETHZ-390382/2020\|EPI_ISL_72 | ···················· | ···················· | ···················· | ···················· | ···················· | ···················· |
| X | 3527 hCoV-19/Switzerland/GR-ETHZ-390383/2020\|EPI_ISL_72 | ···················· | ···················· | ···················· | ···················· | ···················· | ···················· |
| X | 3528 hCoV-19/Switzerland/GR-ETHZ-390389/2020\|EPI_ISL_72 | ···················· | ···················· | ···················· | ···················· | ···················· | ···················· |
| X | 3529 hCoV-19/Switzerland/LU-ETHZ-380331/2020\|EPI_ISL_72 | ···················· | ···················· | ···················· | ···················· | ···················· | ···················· |
| X | 3530 hCoV-19/Switzerland/LU-ETHZ-380333/2020\|EPI_ISL_72 | ···················· | ···················· | ···················· | ···················· | ···················· | ···················· |
| X | 3531 hCoV-19/Switzerland/NE-ETHZ-390319/2020\|EPI_ISL_72 | ···················· | ···················· | ···················· | ···················· | ···················· | ···················· |
| X | 3532 hCoV-19/England/CAMC-B7B2C3/2020\|EPI_ISL_724000\|20 | ···················· | ···················· | ···················· | ···················· | ···················· | ···················· |
| X | 3533 hCoV-19/England/CAMC-B7B2D2/2020\|EPI_ISL_724001\|20 | ···················· | ···················· | ···················· | ···················· | ···················· | ···················· |
| X | 3534 hCoV-19/England/CAMC-B7B2F0/2020\|EPI_ISL_724002\|20 | ···················· | ···················· | ···················· | ···················· | ·····T·············· | ···················· |
| X | 3535 hCoV-19/England/CAMC-B7B30C/2020\|EPI_ISL_724003\|20 | ···················· | ···················· | ···················· | ···················· | ···················· | ···················· |

| | | | | | | | |
|---|---|---|---|---|---|---|---|
| X | 3536 hCoV-19/England/CAMC-B7B32A/2020\|EPI_ISL_724004\|20 | ···················· | ···················· | ····················· | ···················· | ···················· | ···················· |
| X | 3537 hCoV-19/England/CAMC-B7B393/2020\|EPI_ISL_724005\|20 | ···················· | ···················· | ····················· | ···················· | ···················· | ···················· |
| X | 3538 hCoV-19/England/CAMC-B7B3A2/2020\|EPI_ISL_724006\|20 | ···················· | ···················· | ····················· | ···················· | ···················· | ···················· |
| X | 3539 hCoV-19/England/CAMC-B7B472/2020\|EPI_ISL_724011\|20 | ···················· | ···················· | ····················· | ···················· | ···················· | ···················· |
| X | 3540 hCoV-19/England/CAMC-B7B481/2020\|EPI_ISL_724012\|20 | ···················· | ···················· | ····················· | ···················· | ···················· | ···················· |
| X | 3541 hCoV-19/England/CAMC-B7B490/2020\|EPI_ISL_724013\|20 | ···················· | ···················· | ····················· | ···················· | ···················· | ···················· |
| X | 3542 hCoV-19/England/CAMC-B7B4DC/2020\|EPI_ISL_724014\|20 | ···················· | ···················· | ····················· | ···················· | ···················· | ···················· |
| X | 3543 hCoV-19/England/CAMC-B7B4EB/2020\|EPI_ISL_724015\|20 | ···················· | ···················· | ····················· | ···················· | ···················· | ···················· |
| X | 3544 hCoV-19/England/CAMC-B7B603/2020\|EPI_ISL_724016\|20 | ···················· | ···················· | ····················· | ···················· | ···················· | ···················· |
| X | 3545 hCoV-19/England/CAMC-B7B621/2020\|EPI_ISL_724017\|20 | ···················· | ···················· | ····················· | ···················· | ···················· | ···················· |
| X | 3546 hCoV-19/England/CAMC-B7B630/2020\|EPI_ISL_724018\|20 | ···················· | ···················· | ····················· | ···················· | ···················· | ···················· |
| X | 3547 hCoV-19/England/CAMC-B7B64F/2020\|EPI_ISL_724019\|20 | ···················· | ···················· | ····················· | ···················· | ···················· | ···················· |
| X | 3548 hCoV-19/England/CAMC-B7B65E/2020\|EPI_ISL_724020\|20 | ···················· | ···················· | ····················· | ···················· | ···················· | ···················· |
| X | 3549 hCoV-19/England/CAMC-B7B66D/2020\|EPI_ISL_724021\|20 | ···················· | ···················· | ····················· | ···················· | ···················· | ···················· |
| X | 3550 hCoV-19/England/CAMC-B7B68B/2020\|EPI_ISL_724023\|20 | ···················· | ···················· | ····················· | ···················· | ···················· | ···················· |
| X | 3551 hCoV-19/England/CAMC-B7B69A/2020\|EPI_ISL_724024\|20 | ···················· | ···················· | ····················· | ···················· | ···················· | ···················· |
| X | 3552 hCoV-19/England/CAMC-B7B6A9/2020\|EPI_ISL_724025\|20 | ···················· | ···················· | ····················· | ···················· | ···················· | ···················· |
| X | 3553 hCoV-19/England/CAMC-B7B6B8/2020\|EPI_ISL_724026\|20 | ···················· | ···················· | ····················· | ···················· | ···················· | ···················· |
| X | 3554 hCoV-19/England/CAMC-B7B700/2020\|EPI_ISL_724028\|20 | ···················· | ···················· | ····················· | ···················· | ···················· | ···················· |
| X | 3555 hCoV-19/England/CAMC-B7B71F/2020\|EPI_ISL_724029\|20 | ···················· | ···················· | ····················· | ···················· | ···················· | ···················· |
| X | 3556 hCoV-19/England/CAMC-B7B73D/2020\|EPI_ISL_724030\|20 | ···················· | ···················· | ····················· | ···················· | ···················· | ···················· |
| X | 3557 hCoV-19/England/CAMC-B7B779/2020\|EPI_ISL_724031\|20 | ···················· | ···················· | ····················· | ···················· | ······T············· | ···················· |
| X | 3558 hCoV-19/England/CAMC-B7B797/2020\|EPI_ISL_724032\|20 | ···················· | ···················· | ····················· | ···················· | ···················· | ···················· |
| X | 3559 hCoV-19/England/CAMC-B7B7D3/2020\|EPI_ISL_724035\|20 | ···················· | ···················· | ····················· | ···················· | ···················· | ···················· |
| X | 3560 hCoV-19/England/CAMC-B7B7E2/2020\|EPI_ISL_724036\|20 | ···················· | ···················· | ····················· | ···················· | ······T············· | ···················· |
| X | 3561 hCoV-19/England/CAMC-B7B928/2020\|EPI_ISL_724039\|20 | ···················· | ···················· | ····················· | ···················· | ···················· | ···················· |
| X | 3562 hCoV-19/England/CAMC-B7B937/2020\|EPI_ISL_724040\|20 | ···················· | ···················· | ····················· | ···················· | ···················· | ···················· |
| X | 3563 hCoV-19/England/CAMC-B7B964/2020\|EPI_ISL_724041\|20 | ···················· | ···················· | ····················· | ···················· | ···················· | ···················· |
| X | 3564 hCoV-19/England/CAMC-B7B982/2020\|EPI_ISL_724042\|20 | ···················· | ···················· | ····················· | ···················· | ···················· | ···················· |
| X | 3565 hCoV-19/England/CAMC-B7BC3E/2020\|EPI_ISL_724043\|20 | ···················· | ···················· | ····················· | ···················· | ···················· | ···················· |
| X | 3566 hCoV-19/England/CAMC-B7BC4D/2020\|EPI_ISL_724044\|20 | ···················· | ···················· | ····················· | ···················· | ···················· | ···················· |
| X | 3567 hCoV-19/England/CAMC-B7BC7A/2020\|EPI_ISL_724045\|20 | ···················· | ···················· | ····················· | ···················· | ···················· | ···················· |
| X | 3568 hCoV-19/England/CAMC-B7BCB6/2020\|EPI_ISL_724049\|20 | ···················· | ···················· | ····················· | ···················· | ···················· | ···················· |
| X | 3569 hCoV-19/England/CAMC-B7BCC5/2020\|EPI_ISL_724050\|20 | ···················· | ···················· | ····················· | ···················· | ···················· | ···················· |
| X | 3570 hCoV-19/England/CAMC-B7BD1D/2020\|EPI_ISL_724053\|20 | ···················· | ···················· | ····················· | ···················· | ···················· | ···················· |
| X | 3571 hCoV-19/England/CAMC-B7BD3B/2020\|EPI_ISL_724054\|20 | ···················· | ···················· | ····················· | ···················· | ···················· | ···················· |
| X | 3572 hCoV-19/England/CAMC-B7BD68/2020\|EPI_ISL_724055\|20 | ···················· | ···················· | ····················· | ···················· | ···················· | ···················· |
| X | 3573 hCoV-19/England/CAMC-B7BD77/2020\|EPI_ISL_724056\|20 | ···················· | ···················· | ····················· | ···················· | ···················· | ···················· |
| X | 3574 hCoV-19/England/CAMC-B7BD86/2020\|EPI_ISL_724057\|20 | ···················· | ···················· | ····················· | ···················· | ···················· | ···················· |
| X | 3575 hCoV-19/England/CAMC-B7BD95/2020\|EPI_ISL_724058\|20 | ···················· | ···················· | ····················· | ···················· | ···················· | ···················· |
| X | 3576 hCoV-19/England/CAMC-B7BE74/2020\|EPI_ISL_724059\|20 | ···················· | ···················· | ····················· | ···················· | ···················· | ···················· |
| X | 3577 hCoV-19/England/CAMC-B7BE83/2020\|EPI_ISL_724060\|20 | ···················· | ···················· | ····················· | ···················· | ···················· | ···················· |
| X | 3578 hCoV-19/England/CAMC-B7BE92/2020\|EPI_ISL_724061\|20 | ········T··········· | ···················· | ····················· | ···················· | ···················· | ···················· |
| X | 3579 hCoV-19/England/CAMC-B7BEA1/2020\|EPI_ISL_724062\|20 | ···················· | ···················· | ····················· | ···················· | ······T············· | ···················· |
| X | 3580 hCoV-19/England/CAMC-BBB7B9/2020\|EPI_ISL_724067\|20 | ···················· | ···················· | ····················· | ···················· | ···················· | ···················· |
| X | 3581 hCoV-19/England/CAMC-BBBB26/2020\|EPI_ISL_724090\|20 | ···················· | ···················· | ····················· | ···················· | ······T············· | ···················· |
| X | 3582 hCoV-19/England/CAMC-BBBB35/2020\|EPI_ISL_724091\|20 | ···················· | ···················· | ····················· | ···················· | ···················· | ···················· |
| X | 3583 hCoV-19/England/CAMC-BBBB71/2020\|EPI_ISL_724094\|20 | ···················· | ······A············· | ····················· | ···················· | ···················· | ···················· |
| X | 3584 hCoV-19/England/CAMC-BBBBDB/2020\|EPI_ISL_724097\|20 | ···················· | ···················· | ····················· | ···················· | ···················· | ···················· |
| X | 3585 hCoV-19/Denmark/DCGC-16326/2020\|EPI_ISL_714704\|202 | ···················· | ···················· | ····················· | ···················· | ···················· | ···················· |
| X | 3586 hCoV-19/Malaysia/UNIMAS-15792/2020\|EPI_ISL_718200\| | ···················· | ·A·················· | ····················· | ···················· | ···················· | ···················· |
| X | 3587 hCoV-19/Malaysia/UNIMAS-15731/2020\|EPI_ISL_718201\| | ···················· | ·A·················· | ····················· | ···················· | ···················· | ···················· |
| X | 3588 hCoV-19/Malaysia/UNIMAS-15796/2020\|EPI_ISL_718202\| | ···················· | ·A·················· | ····················· | ···················· | ···················· | ···················· |
| X | 3589 hCoV-19/Malaysia/UNIMAS-B054/2020\|EPI_ISL_718203\|2 | ···················· | ·A·················· | ····················· | ···················· | ···················· | ···················· |
| X | 3590 hCoV-19/Malaysia/UNIMAS-B021/2020\|EPI_ISL_718204\|2 | ···················· | ·A·················· | ····················· | ···················· | ···················· | ···················· |
| X | 3591 hCoV-19/Malaysia/UNIMAS-B060/2020\|EPI_ISL_718205\|2 | ···················· | ·A·················· | ····················· | ···················· | ···················· | ···················· |
| X | 3592 hCoV-19/Malaysia/UNIMAS-15718/2020\|EPI_ISL_718206\| | ···················· | ·A·················· | ····················· | ···················· | ···················· | ···················· |
| X | 3593 hCoV-19/Malaysia/UNIMAS-15736/2020\|EPI_ISL_718207\| | ···················· | ·A·················· | ····················· | ···················· | ···················· | ···················· |
| X | 3594 hCoV-19/Malaysia/UNIMAS-15712/2020\|EPI_ISL_718208\| | ···················· | ·A·················· | ····················· | ···················· | ···················· | ···················· |
| X | 3595 hCoV-19/Malaysia/UNIMAS-15714/2020\|EPI_ISL_718209\| | ···················· | ·A·················· | ····················· | ···················· | ···················· | ···················· |
| X | 3596 hCoV-19/Malaysia/UNIMAS-15279/2020\|EPI_ISL_718210\| | ···················· | ·A·················· | ····················· | ···················· | ···················· | ···················· |
| X | 3597 hCoV-19/Malaysia/UNIMAS-15681/2020\|EPI_ISL_718211\| | ···················· | ·A·················· | ····················· | ···················· | ···················· | ···················· |
| X | 3598 hCoV-19/Malaysia/UNIMAS-15683/2020\|EPI_ISL_718212\| | ···················· | ·A·················· | ····················· | ···················· | ···················· | ···················· |
| X | 3599 hCoV-19/Malaysia/UNIMAS-15682/2020\|EPI_ISL_718213\| | ···················· | ·A·················· | ····················· | ···················· | ···················· | ···················· |
| X | 3600 hCoV-19/Malaysia/UNIMAS-15287/2020\|EPI_ISL_718214\| | ···················· | ·A·················· | ····················· | ···················· | ···················· | ···················· |
| X | 3601 hCoV-19/Malaysia/UNIMAS-15288/2020\|EPI_ISL_718215\| | ···················· | ·A·················· | ····················· | ···················· | ···················· | ···················· |
| X | 3602 hCoV-19/Malaysia/UNIMAS-B072/2020\|EPI_ISL_718216\|2 | ···················· | ·A·················· | ····················· | ···················· | ···················· | ···················· |
| X | 3603 hCoV-19/Malaysia/UNIMAS-15710/2020\|EPI_ISL_718217\| | ···················· | ·A·················· | ····················· | ···················· | ···················· | ···················· |
| X | 3604 hCoV-19/Malaysia/UNIMAS-B023/2020\|EPI_ISL_718218\|2 | ···················· | ·A·················· | ····················· | ···················· | ···················· | ···················· |
| X | 3605 hCoV-19/Malaysia/UNIMAS-15784/2020\|EPI_ISL_718220\| | ···················· | ···················· | ····················· | ···················· | ···················· | ···················· |
| X | 3606 hCoV-19/Malaysia/UNIMAS-2426/2020\|EPI_ISL_718221\|2 | ···················· | ···················· | ····················· | ···················· | ···················· | ···················· |
| X | 3607 hCoV-19/Malaysia/UNIMAS-B026/2020\|EPI_ISL_718222\|2 | ···················· | ·A·················· | ····················· | ···················· | ···················· | ···················· |
| X | 3608 hCoV-19/France/BRE-IPP9966/2020\|EPI_ISL_718226\|202 | ···················· | ···················· | ····················· | ···················· | ···················· | ···················· |
| X | 3609 hCoV-19/France/BRE-IPP9967/2020\|EPI_ISL_718227\|202 | ···················· | ···················· | ····················· | ···················· | ···················· | ···················· |
| X | 3610 hCoV-19/France/BRE-IPP9969/2020\|EPI_ISL_718228\|202 | ···················· | ···················· | ····················· | ···················· | ···················· | ···················· |
| X | 3611 hCoV-19/France/BRE-IPP9970/2020\|EPI_ISL_718229\|202 | ···················· | ···················· | ····················· | ···················· | ···················· | ···················· |
| X | 3612 hCoV-19/France/BRE-IPP9971/2020\|EPI_ISL_718230\|202 | ···················· | ···················· | ····················· | ···················· | ······T············· | ···················· |
| X | 3613 hCoV-19/France/BRE-IPP9972/2020\|EPI_ISL_718231\|202 | ···················· | ···················· | ····················· | ···················· | ···················· | ···················· |
| X | 3614 hCoV-19/France/BRE-IPP9973/2020\|EPI_ISL_718232\|202 | ···················· | ···················· | ····················· | ···················· | ···················· | ···················· |
| X | 3615 hCoV-19/France/BRE-IPP9975/2020\|EPI_ISL_718233\|202 | ···················· | ···················· | ····················· | ···················· | ···················· | ···················· |

| | | | | | | | |
|---|---|---|---|---|---|---|---|
| X | 3616 hCoV-19/France/BRE-IPP9976/2020\|EPI_ISL_718234\|202 | ···················· | ···················· | ····················· | ···················· | ···················· | ···················· |
| X | 3617 hCoV-19/France/BRE-IPP9978/2020\|EPI_ISL_718235\|202 | ···················· | ···················· | ····················· | ···················· | ···················· | ···················· |
| X | 3618 hCoV-19/France/BRE-IPP9979/2020\|EPI_ISL_718236\|202 | ···················· | ···················· | ····················· | ···················· | ···················· | ···················· |
| X | 3619 hCoV-19/France/BRE-IPP9980/2020\|EPI_ISL_718237\|202 | ···················· | ···················· | ····················· | ···················· | ···················· | ···················· |
| X | 3620 hCoV-19/France/BRE-IPP9981/2020\|EPI_ISL_718238\|202 | ···················· | ···················· | ····················· | ···················· | ···················· | ···················· |
| X | 3621 hCoV-19/France/BRE-IPP9982/2020\|EPI_ISL_718239\|202 | ···················· | ···················· | ····················· | ···················· | ···················· | ···················· |
| X | 3622 hCoV-19/France/BRE-IPP9983/2020\|EPI_ISL_718240\|202 | ···················· | ···················· | ····················· | ···················· | ···················· | ··············A····· |
| X | 3623 hCoV-19/France/BRE-IPP9984/2020\|EPI_ISL_718241\|202 | ···················· | ···················· | ····················· | ···················· | ···················· | ···················· |
| X | 3624 hCoV-19/France/BRE-IPP9986/2020\|EPI_ISL_718242\|202 | ···················· | ···················· | ····················· | ···················· | ···················· | ···················· |
| X | 3625 hCoV-19/France/BRE-IPP9988/2020\|EPI_ISL_718243\|202 | ···················· | ···················· | ····················· | ···················· | ···················· | ···················· |
| X | 3626 hCoV-19/France/BRE-IPP9989/2020\|EPI_ISL_718244\|202 | ···················· | ···················· | ····················· | ···················· | ···················· | ···················· |
| X | 3627 hCoV-19/France/BRE-IPP9990/2020\|EPI_ISL_718245\|202 | ···················· | ···················· | ····················· | ···················· | ···················· | ···················· |
| X | 3628 hCoV-19/France/BRE-IPP9992/2020\|EPI_ISL_718246\|202 | ···················· | ···················· | ····················· | ···················· | ···················· | ···················· |
| X | 3629 hCoV-19/France/BRE-IPP9994/2020\|EPI_ISL_718247\|202 | ···················· | ···················· | ····················· | ···················· | ···················· | ···················· |
| X | 3630 hCoV-19/France/BRE-IPP9995/2020\|EPI_ISL_718248\|202 | ···················· | ···················· | ····················· | ···················· | ···················· | ···················· |
| X | 3631 hCoV-19/France/BRE-IPP9996/2020\|EPI_ISL_718249\|202 | ···················· | ···················· | ····················· | ···················· | ···················· | ···················· |
| X | 3632 hCoV-19/Slovakia/UKBA-402/2020\|EPI_ISL_718250\|2020 | ···················· | ···················· | ····················· | ···················· | ···················· | ···················· |
| X | 3633 hCoV-19/Slovakia/UKBA-403/2020\|EPI_ISL_718251\|2020 | ···················· | ···················· | ····················· | ···················· | ···················· | ···················· |
| X | 3634 hCoV-19/Slovakia/UKBA-404/2020\|EPI_ISL_718252\|2020 | ···················· | ···················· | ····················· | ···················· | ···················· | ···················· |
| X | 3635 hCoV-19/Slovakia/UKBA-405/2020\|EPI_ISL_718253\|2020 | ···················· | ···················· | ····················· | ···················· | ···················· | ···················· |
| X | 3636 hCoV-19/Slovakia/UKBA-406/2020\|EPI_ISL_718254\|2020 | ···················· | ···················· | ····················· | ···················· | ···················· | ···················· |
| X | 3637 hCoV-19/Slovakia/UKBA-407/2020\|EPI_ISL_718255\|2020 | ···················· | ···················· | ····················· | ···················· | ···················· | ···················· |
| X | 3638 hCoV-19/Slovakia/UKBA-408/2020\|EPI_ISL_718256\|2020 | ···················· | ···················· | ····················· | ···················· | ···················· | ···················· |
| X | 3639 hCoV-19/Slovakia/UKBA-409/2020\|EPI_ISL_718257\|2020 | ···················· | ···················· | ····················· | ···················· | ···················· | ···················· |
| X | 3640 hCoV-19/Slovakia/UKBA-410/2020\|EPI_ISL_718258\|2020 | ···················· | ···················· | ····················· | ···················· | ···················· | ···················· |
| X | 3641 hCoV-19/Slovakia/UKBA-411/2020\|EPI_ISL_718259\|2020 | ···················· | ···················· | ····················· | ···················· | ···················· | ···················· |
| X | 3642 hCoV-19/Slovakia/UKBA-412/2020\|EPI_ISL_718260\|2020 | ···················· | ···················· | ····················· | ···················· | ···················· | ···················· |
| X | 3643 hCoV-19/Italy/LAZ-INMI-97/2020\|EPI_ISL_718262\|2020 | ···················· | ···················· | ····················· | ···················· | ···················· | ···················· |
| X | 3644 hCoV-19/Italy/LAZ-INMI-98/2020\|EPI_ISL_718263\|2020 | ···················· | ···················· | ····················· | ···················· | ···················· | ···················· |
| X | 3645 hCoV-19/Italy/LAZ-INMI-99/2020\|EPI_ISL_718264\|2020 | ···················· | ···················· | ····················· | ···················· | ···················· | ···················· |
| X | 3646 hCoV-19/Italy/LAZ-INMI-100/2020\|EPI_ISL_718265\|202 | ···················· | ···················· | ····················· | ···················· | ·····T·············· | ···················· |
| X | 3647 hCoV-19/Malaysia/IMR-WI080/2020\|EPI_ISL_718267\|202 | ···················· | ···················· | ····················· | ···················· | ···················· | ···················· |
| X | 3648 hCoV-19/Malaysia/IMR-WI093/2020\|EPI_ISL_718268\|202 | ···················· | ···················· | ····················· | ···················· | ···················· | ···················· |
| X | 3649 hCoV-19/Malaysia/IMR-CV138548/2020\|EPI_ISL_718290\| | ···················· | ·A·················· | ····················· | ···················· | ···················· | ···················· |
| X | 3650 hCoV-19/Malaysia/IMR-CV138551/2020\|EPI_ISL_718291\| | ···················· | ·A·················· | ····················· | ···················· | ···················· | ···················· |
| X | 3651 hCoV-19/Malaysia/IMR-CV138687/2020\|EPI_ISL_718292\| | ···················· | ·A·················· | ····················· | ···················· | ···················· | ···················· |
| X | 3652 hCoV-19/Malaysia/IMR-CV138815/2020\|EPI_ISL_718293\| | ···················· | ·A·················· | ····················· | ···················· | ···················· | ···················· |
| X | 3653 hCoV-19/Malaysia/IMR-CV139140/2020\|EPI_ISL_718294\| | ···················· | ·A·················· | ····················· | ···················· | ···················· | ···················· |
| X | 3654 hCoV-19/Malaysia/IMR-CV139158/2020\|EPI_ISL_718296\| | ···················· | ·A·················· | ····················· | ···················· | ···················· | ···················· |
| X | 3655 hCoV-19/Malaysia/IMR-CV139176/2020\|EPI_ISL_718297\| | ···················· | ·A·················· | ····················· | ···················· | ···················· | ···················· |
| X | 3656 hCoV-19/Malaysia/IMR-CV139206/2020\|EPI_ISL_718298\| | ···················· | ·A·················· | ····················· | ···················· | ···················· | ···················· |
| X | 3657 hCoV-19/Malaysia/IMR-CV1140107/2020\|EPI_ISL_718299 | ···················· | ·A·················· | ····················· | ···················· | ···················· | ···················· |
| X | 3658 hCoV-19/England/CAMC-C44435/2020\|EPI_ISL_719518\|20 | ···················· | ···················· | ····················· | ···················· | ···················· | ···················· |
| X | 3659 hCoV-19/England/CAMC-C44945/2020\|EPI_ISL_719523\|20 | ···················· | ···················· | ····················· | ···················· | ···················· | ···················· |
| X | 3660 hCoV-19/England/CAMC-C4580B/2020\|EPI_ISL_719538\|20 | ···················· | ···················· | ····················· | ···················· | ···················· | ···················· |
| X | 3661 hCoV-19/England/CAMC-C4406E/2020\|EPI_ISL_719540\|20 | ···················· | ···················· | ····················· | ···················· | ···················· | ···················· |
| X | 3662 hCoV-19/England/CAMC-C458A1/2020\|EPI_ISL_719545\|20 | ···················· | ···················· | ····················· | ···················· | ···················· | ···················· |
| X | 3663 hCoV-19/England/CAMC-C3F3A9/2020\|EPI_ISL_719548\|20 | ···················· | ···················· | ····················· | ···················· | ···················· | ···················· |
| X | 3664 hCoV-19/England/CAMC-C441A7/2020\|EPI_ISL_719551\|20 | ···················· | ···················· | ····················· | ···················· | ···················· | ···················· |
| X | 3665 hCoV-19/England/CAMC-C4564D/2020\|EPI_ISL_719564\|20 | ···················· | ···················· | ····················· | ···················· | ···················· | ···················· |
| X | 3666 hCoV-19/England/CAMC-C4390A/2020\|EPI_ISL_719566\|20 | ···················· | ···················· | ····················· | ···················· | ···················· | ···················· |
| X | 3667 hCoV-19/England/CAMC-C441D4/2020\|EPI_ISL_719570\|20 | ···················· | ···················· | ····················· | ···················· | ···················· | ···················· |
| X | 3668 hCoV-19/England/CAMC-C3F5EF/2020\|EPI_ISL_719571\|20 | ···················· | ···················· | ····················· | ···················· | ···················· | ···················· |
| X | 3669 hCoV-19/England/CAMC-C433C0/2020\|EPI_ISL_719574\|20 | ···················· | ···················· | ····················· | ···················· | ···················· | ···················· |
| X | 3670 hCoV-19/England/CAMC-C443DE/2020\|EPI_ISL_719576\|20 | ···················· | ···················· | ····················· | ···················· | ···················· | ···················· |
| X | 3671 hCoV-19/England/CAMC-C43F53/2020\|EPI_ISL_719578\|20 | ···················· | ·A·················· | ····················· | ···················· | ···················· | ··A················· |
| X | 3672 hCoV-19/England/CAMC-C43E47/2020\|EPI_ISL_719581\|20 | ···················· | ···················· | ····················· | ···················· | ·····T·············· | ···················· |
| X | 3673 hCoV-19/England/CAMC-C42473/2020\|EPI_ISL_719583\|20 | ···················· | ···················· | ····················· | ···················· | ···················· | ···················· |
| X | 3674 hCoV-19/England/CAMC-C45276/2020\|EPI_ISL_719596\|20 | ···················· | ···················· | ····················· | ···················· | ···················· | ···················· |
| X | 3675 hCoV-19/England/MILK-BB0A24/2020\|EPI_ISL_720504\|20 | ···················· | ···················· | ····················· | ···················· | ···················· | ···················· |
| X | 3676 hCoV-19/England/MILK-BB1B4E/2020\|EPI_ISL_720506\|20 | ···················· | ···················· | ····················· | ···················· | ···················· | ···················· |
| X | 3677 hCoV-19/England/MILK-BB0DD0/2020\|EPI_ISL_720507\|20 | ···················· | ···················· | ····················· | ···················· | ···················· | ···················· |
| X | 3678 hCoV-19/England/MILK-BB1AAB/2020\|EPI_ISL_720508\|20 | ···················· | ·A·················· | ····················· | ···················· | ···················· | ··A················· |
| X | 3679 hCoV-19/England/MILK-BB1DB1/2020\|EPI_ISL_720509\|20 | ···················· | ···················· | ····················· | ···················· | ···················· | ···················· |
| X | 3680 hCoV-19/England/MILK-BAC3E4/2020\|EPI_ISL_720510\|20 | ···················· | ···················· | ····················· | ···················· | ···················· | ···················· |
| X | 3681 hCoV-19/England/MILK-BAC812/2020\|EPI_ISL_720511\|20 | ···················· | ·A·················· | ····················· | ···················· | ···················· | ··A················· |
| X | 3682 hCoV-19/England/MILK-BB09DC/2020\|EPI_ISL_720512\|20 | ···················· | ·A·················· | ····················· | ···················· | ···················· | ··A················· |
| X | 3683 hCoV-19/England/MILK-BAFBE9/2020\|EPI_ISL_720514\|20 | ···················· | ···················· | ····················· | ···················· | ···················· | ···················· |
| X | 3684 hCoV-19/England/MILK-BB0E91/2020\|EPI_ISL_720515\|20 | ···················· | ···················· | ····················· | ···················· | ···················· | ···················· |
| X | 3685 hCoV-19/England/MILK-BB0DFE/2020\|EPI_ISL_720517\|20 | ···················· | ···················· | ····················· | ···················· | ···················· | ···················· |
| X | 3686 hCoV-19/England/MILK-BABA2C/2020\|EPI_ISL_720518\|20 | ···················· | ···················· | ····················· | ···················· | ···················· | ···················· |
| X | 3687 hCoV-19/England/MILK-BB10D6/2020\|EPI_ISL_720519\|20 | ···················· | ···················· | ····················· | ···················· | ···················· | ···················· |
| X | 3688 hCoV-19/England/MILK-BABF4B/2020\|EPI_ISL_720520\|20 | ···················· | ···················· | ····················· | ···················· | ···················· | ···················· |
| X | 3689 hCoV-19/England/MILK-BAB8AA/2020\|EPI_ISL_720521\|20 | ···················· | ···················· | ····················· | ···················· | ···················· | ···················· |
| X | 3690 hCoV-19/England/MILK-BB0BA9/2020\|EPI_ISL_720522\|20 | ···················· | ···················· | ····················· | ···················· | ···················· | ···················· |
| X | 3691 hCoV-19/England/MILK-BB1A32/2020\|EPI_ISL_720523\|20 | ···················· | ···················· | ····················· | ···················· | ···················· | ···················· |
| X | 3692 hCoV-19/England/MILK-BB0787/2020\|EPI_ISL_720524\|20 | ···················· | ···················· | ····················· | ···················· | ···················· | ···················· |
| X | 3693 hCoV-19/England/MILK-BB114C/2020\|EPI_ISL_720525\|20 | ···················· | ···················· | ····················· | ···················· | ···················· | ···················· |
| X | 3694 hCoV-19/England/MILK-BAB87D/2020\|EPI_ISL_720527\|20 | ···················· | ···················· | ····················· | ···················· | ···················· | ···················· |
| X | 3695 hCoV-19/England/MILK-BB2345/2020\|EPI_ISL_720528\|20 | ···················· | ···················· | ····················· | ···················· | ···················· | ···················· |

X 3696 hCoV-19/England/MILK-BB1A14/2020|EPI_ISL_720530|20 ···················· ···················· ····················· ···················· ···················· ····················
X 3697 hCoV-19/England/MILK-BB0B03/2020|EPI_ISL_720531|20 ···················· ···················· ····················· ···················· ···················· ····················
X 3698 hCoV-19/England/MILK-BAFAEC/2020|EPI_ISL_720532|20 ···················· ···················· ····················· ···················· ···················· ····················
X 3699 hCoV-19/England/MILK-BAFA91/2020|EPI_ISL_720533|20 ···················· ···················· ····················· ···················· ···················· ····················
X 3700 hCoV-19/England/MILK-BB0E37/2020|EPI_ISL_720534|20 ···················· ···················· ····················· ···················· ···················· ····················
X 3701 hCoV-19/England/MILK-BB21C3/2020|EPI_ISL_720535|20 ···················· ···················· ····················· ···················· ···················· ····················
X 3702 hCoV-19/England/MILK-BB1A50/2020|EPI_ISL_720536|20 ···················· ···················· ····················· ···················· ···················· ····················
X 3703 hCoV-19/England/MILK-BB03DE/2020|EPI_ISL_720537|20 ···················· ···················· ····················· ···················· ···················· ····················
X 3704 hCoV-19/England/MILK-BAC539/2020|EPI_ISL_720538|20 ···················· ···················· ····················· ···················· ···················· ····················
X 3705 hCoV-19/England/MILK-BB1D66/2020|EPI_ISL_720540|20 ···················· ···················· ····················· ···················· ···················· ····················
X 3706 hCoV-19/England/MILK-BB21E1/2020|EPI_ISL_720542|20 ···················· ···················· ····················· ···················· ···················· ····················
X 3707 hCoV-19/England/MILK-BB0B7C/2020|EPI_ISL_720545|20 ···················· ···················· ····················· ···················· ···················· ····················
X 3708 hCoV-19/England/MILK-BABFE1/2020|EPI_ISL_720546|20 ···················· ···················· ····················· ···················· ···················· ····················
X 3709 hCoV-19/England/MILK-BB0AD9/2020|EPI_ISL_720547|20 ···················· ···················· ····················· ···················· ···················· ····················
X 3710 hCoV-19/England/MILK-BB06E4/2020|EPI_ISL_720548|20 ···················· ···················· ····················· ···················· ···················· ····················
X 3711 hCoV-19/England/MILK-BAC37B/2020|EPI_ISL_720549|20 ···················· ···················· ····················· ···················· ···················· ····················
X 3712 hCoV-19/England/MILK-BB0347/2020|EPI_ISL_720550|20 ···················· ·A·················· ····················· ···················· ···················· ··A·················
X 3713 hCoV-19/England/MILK-BB1B5D/2020|EPI_ISL_720552|20 ···················· ···················· ····················· ···················· ···················· ····················
X 3714 hCoV-19/England/MILK-BB0004/2020|EPI_ISL_720553|20 ···················· ···················· ····················· ···················· ···················· ····················
X 3715 hCoV-19/England/MILK-BB07E1/2020|EPI_ISL_720554|20 ···················· ···················· ····················· ···················· ···················· ····················
X 3716 hCoV-19/England/MILK-BB2011/2020|EPI_ISL_720555|20 ···················· ···················· ····················· ···················· ···················· ····················
X 3717 hCoV-19/England/MILK-BABA77/2020|EPI_ISL_720556|20 ···················· ···················· ····················· ···················· ···················· ····················
X 3718 hCoV-19/England/MILK-BABAB3/2020|EPI_ISL_720557|20 ···················· ···················· ····················· ···················· ···················· ····················
X 3719 hCoV-19/England/MILK-BB107C/2020|EPI_ISL_720558|20 ···················· ···················· ····················· ···················· ···················· ····················
X 3720 hCoV-19/England/MILK-BAC3B7/2020|EPI_ISL_720560|20 ···················· ···················· ····················· ···················· ···················· ····················
X 3721 hCoV-19/England/MILK-BB0EEC/2020|EPI_ISL_720561|20 ···················· ···················· ····················· ···················· ···················· ····················
X 3722 hCoV-19/England/MILK-BB009B/2020|EPI_ISL_720562|20 ···················· ···················· ····················· ···················· ······T············· ····················
X 3723 hCoV-19/England/MILK-BAC302/2020|EPI_ISL_720564|20 ···················· ···················· ····················· ···················· ···················· ····················
X 3724 hCoV-19/England/MILK-BB1100/2020|EPI_ISL_720565|20 ···················· ···················· ····················· ···················· ···················· ····················
X 3725 hCoV-19/England/MILK-BB0B5E/2020|EPI_ISL_720567|20 ···················· ···················· ····················· ···················· ···················· ····················
X 3726 hCoV-19/England/MILK-BB006E/2020|EPI_ISL_720568|20 ···················· ···················· ····················· ···················· ···················· ····················
X 3727 hCoV-19/England/MILK-BABA68/2020|EPI_ISL_720569|20 ···················· ···················· ····················· ···················· ···················· ····················
X 3728 hCoV-19/England/MILK-BAC584/2020|EPI_ISL_720570|20 ···················· ···················· ····················· ···················· ···················· ····················
X 3729 hCoV-19/England/MILK-BB0963/2020|EPI_ISL_720571|20 ···················· ·A·················· ····················· ···················· ···················· ··A·················
X 3730 hCoV-19/England/MILK-BB0C2E/2020|EPI_ISL_720572|20 ···················· ···················· ····················· ···················· ···················· ····················
X 3731 hCoV-19/England/MILK-BB1D2A/2020|EPI_ISL_720573|20 ···················· ···················· ····················· ···················· ···················· ····················
X 3732 hCoV-19/England/MILK-BB0A33/2020|EPI_ISL_720574|20 ···················· ···················· ····················· ···················· ···················· ····················
X 3733 hCoV-19/England/MILK-BB0DB2/2020|EPI_ISL_720575|20 ···················· ···················· ····················· ···················· ···················· ····················
X 3734 hCoV-19/England/MILK-BAFACE/2020|EPI_ISL_720577|20 ···················· ···················· ····················· ···················· ···················· ····················
X 3735 hCoV-19/England/MILK-BB0295/2020|EPI_ISL_720578|20 ···················· ·A·················· ····················· ···················· ···················· ··A·················
X 3736 hCoV-19/England/MILK-BAFCAA/2020|EPI_ISL_720579|20 ···················· ···················· ····················· ···················· ···················· ····················
X 3737 hCoV-19/England/MILK-BAFD01/2020|EPI_ISL_720581|20 ···················· ···················· ····················· ···················· ···················· ····················
X 3738 hCoV-19/England/MILK-BB0E46/2020|EPI_ISL_720582|20 ···················· ···················· ····················· ···················· ···················· ····················
X 3739 hCoV-19/England/MILK-BB00AA/2020|EPI_ISL_720583|20 ···················· ···················· ····················· ···················· ···················· ····················
X 3740 hCoV-19/England/MILK-BAFEC2/2020|EPI_ISL_720584|20 ···················· ···················· ····················· ···················· ···················· ····················
X 3741 hCoV-19/England/MILK-BB07A5/2020|EPI_ISL_720585|20 ···················· ···················· ····················· ···················· ···················· ····················
X 3742 hCoV-19/England/MILK-BB1B02/2020|EPI_ISL_720586|20 ···················· ···················· ····················· ···················· ···················· ····················
X 3743 hCoV-19/England/MILK-BAC575/2020|EPI_ISL_720587|20 ···················· ···················· ····················· ···················· ···················· ····················
X 3744 hCoV-19/England/MILK-BB1A8D/2020|EPI_ISL_720589|20 ···················· ···················· ····················· ···················· ···················· ····················
X 3745 hCoV-19/Switzerland/BL-ETHZ-380395/2020|EPI_ISL_72 ···················· ···················· ····················· ···················· ···················· ····················
X 3746 hCoV-19/Switzerland/BS-ETHZ-380231/2020|EPI_ISL_72 ···················· ···················· ····················· ···················· ···················· ····················
X 3747 hCoV-19/Switzerland/BS-ETHZ-380235/2020|EPI_ISL_72 ···················· ···················· ····················· ···················· ···················· ····················
X 3748 hCoV-19/Switzerland/BS-ETHZ-380347/2020|EPI_ISL_72 ···················· ···················· ····················· ···················· ···················· ····················
X 3749 hCoV-19/Switzerland/BS-ETHZ-380385/2020|EPI_ISL_72 ···················· ···················· ····················· ···················· ···················· ····················
X 3750 hCoV-19/Switzerland/BS-ETHZ-380388/2020|EPI_ISL_72 ···················· ···················· ····················· ···················· ···················· ····················
X 3751 hCoV-19/Switzerland/GE-ETHZ-340089/2020|EPI_ISL_72 ···················· ···················· ····················· ···················· ···················· ····················
X 3752 hCoV-19/Switzerland/GE-ETHZ-340094/2020|EPI_ISL_72 ···················· ···················· ····················· ···················· ···················· ····················
X 3753 hCoV-19/Switzerland/GE-ETHZ-340096/2020|EPI_ISL_72 ···················· ···················· ····················· ···················· ······K············· ····················
X 3754 hCoV-19/Switzerland/GE-ETHZ-340099/2020|EPI_ISL_72 ···················· ···················· ····················· ···················· ···················· ····················
X 3755 hCoV-19/Switzerland/GE-ETHZ-340103/2020|EPI_ISL_72 ···················· ···················· ····················· ···················· ···················· ····················
X 3756 hCoV-19/Switzerland/GR-ETHZ-390390/2020|EPI_ISL_72 ···················· ···················· ····················· ···················· ···················· ····················
X 3757 hCoV-19/Switzerland/LU-ETHZ-340087/2020|EPI_ISL_72 ···················· ···················· ····················· ···················· ···················· ····················
X 3758 hCoV-19/Switzerland/LU-ETHZ-340101/2020|EPI_ISL_72 ···················· ···················· ····················· ···················· ···················· ····················
X 3759 hCoV-19/Switzerland/LU-ETHZ-340112/2020|EPI_ISL_72 ···················· ···················· ····················· ···················· ···················· ····················
X 3760 hCoV-19/Switzerland/LU-ETHZ-380230/2020|EPI_ISL_72 ···················· ···················· ····················· ···················· ···················· ····················
X 3761 hCoV-19/Switzerland/LU-ETHZ-380327/2020|EPI_ISL_72 ···················· ···················· ····················· ···················· ···················· ····················
X 3762 hCoV-19/Switzerland/LU-ETHZ-380328/2020|EPI_ISL_72 ···················· ···················· ····················· ···················· ···················· ····················
X 3763 hCoV-19/Switzerland/LU-ETHZ-380329/2020|EPI_ISL_72 ···················· ···················· ····················· ···················· ···················· ····················
X 3764 hCoV-19/Switzerland/NE-ETHZ-390320/2020|EPI_ISL_72 ···················· ···················· ····················· ···················· ···················· ····················
X 3765 hCoV-19/Switzerland/NE-ETHZ-390321/2020|EPI_ISL_72 ···················· ···················· ····················· ···················· ···················· ····················
X 3766 hCoV-19/Switzerland/NE-ETHZ-390322/2020|EPI_ISL_72 ···················· ···················· ····················· ···················· ···················· ····················
X 3767 hCoV-19/Switzerland/NE-ETHZ-390323/2020|EPI_ISL_72 ···················· ···················· ····················· ···················· ···················· ····················
X 3768 hCoV-19/Switzerland/NE-ETHZ-390325/2020|EPI_ISL_72 ···················· ···················· ····················· ···················· ···················· ····················
X 3769 hCoV-19/Switzerland/NE-ETHZ-390327/2020|EPI_ISL_72 ···················· ···················· ····················· ···················· ···················· ····················
X 3770 hCoV-19/Switzerland/NE-ETHZ-390329/2020|EPI_ISL_72 ···················· ···················· ····················· ···················· ···················· ····················
X 3771 hCoV-19/Switzerland/NE-ETHZ-390330/2020|EPI_ISL_72 ···················· ···················· ····················· ···················· ···················· ····················
X 3772 hCoV-19/Switzerland/NE-ETHZ-390331/2020|EPI_ISL_72 ···················· ···················· ····················· ···················· ···················· ····················
X 3773 hCoV-19/Switzerland/NE-ETHZ-390332/2020|EPI_ISL_72 ···················· ···················· ····················· ···················· ···················· ····················
X 3774 hCoV-19/Switzerland/SG-ETHZ-380302/2020|EPI_ISL_72 ···················· ···················· ····················· ···················· ···················· ····················
X 3775 hCoV-19/Switzerland/SG-ETHZ-380372/2020|EPI_ISL_72 ···················· ···················· ····················· ···················· ···················· ····················

X 3776 hCoV-19/Switzerland/SG-ETHZ-380398/2020|EPI_ISL_72 ···················· ···················· ····················· ···················· ···················· ····················
X 3777 hCoV-19/Switzerland/SG-ETHZ-380399/2020|EPI_ISL_72 ···················· ···················· ····················· ···················· ···················· ····················
X 3778 hCoV-19/Switzerland/SG-ETHZ-380401/2020|EPI_ISL_72 ···················· ···················· ····················· ···················· ···················· ····················
X 3779 hCoV-19/Switzerland/SG-ETHZ-390217/2020|EPI_ISL_72 ···················· ···················· ····················· ···················· ···················· ····················
X 3780 hCoV-19/Switzerland/SG-ETHZ-390220/2020|EPI_ISL_72 ···················· ···················· ····················· ···················· ···················· ····················
X 3781 hCoV-19/Switzerland/SG-ETHZ-390221/2020|EPI_ISL_72 ···················· ···················· ····················· ···················· ···················· ····················
X 3782 hCoV-19/Switzerland/SG-ETHZ-390384/2020|EPI_ISL_72 ···················· ·A·················· ····················· ···················· ···················· ····················
X 3783 hCoV-19/Switzerland/SG-ETHZ-390385/2020|EPI_ISL_72 ···················· ···················· ····················· ···················· ···················· ····················
X 3784 hCoV-19/Switzerland/SG-ETHZ-390386/2020|EPI_ISL_72 ···················· ···················· ····················· ···················· ···················· ····················
X 3785 hCoV-19/Switzerland/SH-ETHZ-340108/2020|EPI_ISL_72 ···················· ···················· ····················· ···················· ···················· ····················
X 3786 hCoV-19/Switzerland/SH-ETHZ-340109/2020|EPI_ISL_72 ···················· ···················· ····················· ···················· ···················· ····················
X 3787 hCoV-19/Switzerland/SH-ETHZ-340114/2020|EPI_ISL_72 ···················· ···················· ····················· ···················· ···················· ····················
X 3788 hCoV-19/Switzerland/SH-ETHZ-340166/2020|EPI_ISL_72 ···················· ·A·················· ····················· ···················· ···················· ···A················
X 3789 hCoV-19/Switzerland/SH-ETHZ-380203/2020|EPI_ISL_72 ···················· ···················· ····················· ···················· ···················· ····················
X 3790 hCoV-19/Switzerland/SH-ETHZ-380204/2020|EPI_ISL_72 ···················· ···················· ····················· ···················· ···················· ····················
X 3791 hCoV-19/Switzerland/SH-ETHZ-380211/2020|EPI_ISL_72 ···················· ·A·················· ····················· ···················· ···················· ····················
X 3792 hCoV-19/Switzerland/SH-ETHZ-380216/2020|EPI_ISL_72 ···················· ···················· ····················· ···················· ···················· ····················
X 3793 hCoV-19/Switzerland/SH-ETHZ-380217/2020|EPI_ISL_72 ···················· ···················· ····················· ···················· ···················· ····················
X 3794 hCoV-19/Switzerland/SH-ETHZ-380219/2020|EPI_ISL_72 ···················· ···················· ····················· ···················· ···················· ····················
X 3795 hCoV-19/Switzerland/SH-ETHZ-380221/2020|EPI_ISL_72 ···················· ···················· ····················· ···················· ···················· ····················
X 3796 hCoV-19/Switzerland/SH-ETHZ-380303/2020|EPI_ISL_72 ···················· ···················· ····················· ···················· ···················· ····················
X 3797 hCoV-19/Switzerland/SH-ETHZ-380309/2020|EPI_ISL_72 ···················· ···················· ····················· ···················· ···················· ····················
X 3798 hCoV-19/Switzerland/SO-ETHZ-340083/2020|EPI_ISL_72 ···················· ···················· ····················· ···················· ···················· ····················
X 3799 hCoV-19/Switzerland/SO-ETHZ-340102/2020|EPI_ISL_72 ···················· ···················· ····················· ···················· ···················· ····················
X 3800 hCoV-19/Switzerland/SO-ETHZ-380237/2020|EPI_ISL_72 ···················· ···················· ····················· ···················· ···················· ····················
X 3801 hCoV-19/Switzerland/SO-ETHZ-380239/2020|EPI_ISL_72 ···················· ···················· ····················· ···················· ···················· ····················
X 3802 hCoV-19/Switzerland/SO-ETHZ-380241/2020|EPI_ISL_72 ···················· ···················· ····················· ···················· ···················· ····················
X 3803 hCoV-19/Switzerland/SO-ETHZ-380243/2020|EPI_ISL_72 ···················· ···················· ····················· ···················· ···················· ····················
X 3804 hCoV-19/Switzerland/SO-ETHZ-380244/2020|EPI_ISL_72 ···················· ···················· ····················· ···················· ···················· ····················
X 3805 hCoV-19/Switzerland/SO-ETHZ-380245/2020|EPI_ISL_72 ···················· ···················· ····················· ···················· ···················· ····················
X 3806 hCoV-19/Switzerland/SO-ETHZ-380279/2020|EPI_ISL_72 ···················· ···················· ····················· ···················· ···················· ····················
X 3807 hCoV-19/Switzerland/SO-ETHZ-380285/2020|EPI_ISL_72 ···················· ···················· ····················· ···················· ···················· ····················
X 3808 hCoV-19/Switzerland/SO-ETHZ-380286/2020|EPI_ISL_72 ···················· ···················· ····················· ···················· ···················· ····················
X 3809 hCoV-19/Switzerland/SO-ETHZ-380289/2020|EPI_ISL_72 ···················· ···················· ····················· ···················· ···················· ····················
X 3810 hCoV-19/Switzerland/SO-ETHZ-380290/2020|EPI_ISL_72 ···················· ···················· ····················· ···················· ···················· ····················
X 3811 hCoV-19/Switzerland/SO-ETHZ-380295/2020|EPI_ISL_72 ···················· ···················· ····················· ···················· ···················· ····················
X 3812 hCoV-19/Switzerland/SO-ETHZ-380297/2020|EPI_ISL_72 ···················· ···················· ····················· ···················· ···················· ····················
X 3813 hCoV-19/Switzerland/SO-ETHZ-380298/2020|EPI_ISL_72 ···················· ···················· ····················· ···················· ······K············· ····················
X 3814 hCoV-19/Switzerland/SO-ETHZ-380301/2020|EPI_ISL_72 ···················· ···················· ····················· ···················· ···················· ····················
X 3815 hCoV-19/Switzerland/SO-ETHZ-380336/2020|EPI_ISL_72 ···················· ···················· ····················· ···················· ···················· ····················
X 3816 hCoV-19/Switzerland/SO-ETHZ-380340/2020|EPI_ISL_72 ···················· ···················· ····················· ···················· ···················· ····················
X 3817 hCoV-19/Switzerland/SO-ETHZ-380351/2020|EPI_ISL_72 ···················· ···················· ····················· ···················· ···················· ····················
X 3818 hCoV-19/Switzerland/SO-ETHZ-380356/2020|EPI_ISL_72 ···················· ·A·················· ····················· ···················· ···················· ···A················
X 3819 hCoV-19/Switzerland/SO-ETHZ-390229/2020|EPI_ISL_72 ···················· ·A·················· ····················· ···················· ···················· ····················
X 3820 hCoV-19/Switzerland/SO-ETHZ-390333/2020|EPI_ISL_72 ···················· ···················· ····················· ···················· ···················· ····················
X 3821 hCoV-19/Switzerland/SO-ETHZ-390334/2020|EPI_ISL_72 ···················· ···················· ····················· ···················· ···················· ····················
X 3822 hCoV-19/Switzerland/SO-ETHZ-390335/2020|EPI_ISL_72 ···················· ···················· ····················· ···················· ···················· ····················
X 3823 hCoV-19/Switzerland/SO-ETHZ-390336/2020|EPI_ISL_72 ···················· ···················· ····················· ···················· ···················· ········R···········
X 3824 hCoV-19/Switzerland/SO-ETHZ-390339/2020|EPI_ISL_72 ···················· ···················· ····················· ···················· ···················· ····················
X 3825 hCoV-19/Switzerland/SO-ETHZ-390340/2020|EPI_ISL_72 ···················· ···················· ····················· ···················· ···················· ····················
X 3826 hCoV-19/Switzerland/SO-ETHZ-390341/2020|EPI_ISL_72 ···················· ···················· ····················· ···················· ···················· ····················
X 3827 hCoV-19/Switzerland/SO-ETHZ-390342/2020|EPI_ISL_72 ···················· ···················· ····················· ···················· ···················· ····················
X 3828 hCoV-19/Switzerland/SZ-ETHZ-340088/2020|EPI_ISL_72 ···················· ·A·················· ····················· ···················· ···················· ····················
X 3829 hCoV-19/Switzerland/SZ-ETHZ-340119/2020|EPI_ISL_72 ···················· ···················· ····················· ···················· ···················· ····················
X 3830 hCoV-19/Switzerland/SZ-ETHZ-340126/2020|EPI_ISL_72 ···················· ···················· ····················· ···················· ···················· ····················
X 3831 hCoV-19/England/CAMC-BBB977/2020|EPI_ISL_724073|20 ···················· ···················· ····················· ···················· ···················· ····················
X 3832 hCoV-19/England/CAMC-BBB986/2020|EPI_ISL_724074|20 ···················· ···················· ····················· ···················· ···················· ····················
X 3833 hCoV-19/England/CAMC-BBB995/2020|EPI_ISL_724075|20 ···················· ···················· ····················· ···················· ···················· ····················
X 3834 hCoV-19/England/CAMC-BBB9A4/2020|EPI_ISL_724076|20 ···················· ···················· ····················· ···················· ···················· ····················
X 3835 hCoV-19/England/CAMC-BBB9FF/2020|EPI_ISL_724078|20 ···················· ···················· ····················· ···················· ···················· ····················
X 3836 hCoV-19/England/CAMC-BBBA0B/2020|EPI_ISL_724079|20 ···················· ···················· ····················· ···················· ···················· ····················
X 3837 hCoV-19/England/CAMC-BBBA65/2020|EPI_ISL_724083|20 ···················· ···················· ····················· ···················· ···················· ····················
X 3838 hCoV-19/England/CAMC-BBBAB0/2020|EPI_ISL_724086|20 ···················· ···················· ····················· ···················· ···················· ····················
X 3839 hCoV-19/England/CAMC-BBBADE/2020|EPI_ISL_724088|20 ···················· ···················· ····················· ···················· ···················· ····················
X 3840 hCoV-19/England/CAMC-BBBB17/2020|EPI_ISL_724089|20 ···················· ···················· ····················· ···················· ······T············· ····················
X 3841 hCoV-19/Denmark/DCGC-14331/2020|EPI_ISL_711700|202 ···················· ···················· ····················· ···················· ···················· ····················
X 3842 hCoV-19/Denmark/DCGC-14336/2020|EPI_ISL_711701|202 ···················· ·A·················· ····················· ···················· ···················· ····················
X 3843 hCoV-19/Denmark/DCGC-14338/2020|EPI_ISL_711702|202 ···················· ···················· ····················· ···················· ···················· ····················
X 3844 hCoV-19/Denmark/DCGC-14339/2020|EPI_ISL_711703|202 ···················· ···················· ····················· ···················· ···················· ····················
X 3845 hCoV-19/Denmark/DCGC-14341/2020|EPI_ISL_711704|202 ···················· ···················· ····················· ···················· ···················· ····················
X 3846 hCoV-19/Denmark/DCGC-14342/2020|EPI_ISL_711705|202 ···················· ···················· ····················· ···················· ···················· ····················
X 3847 hCoV-19/Denmark/DCGC-14343/2020|EPI_ISL_711706|202 ···················· ···················· ····················· ···················· ···················· ····················
X 3848 hCoV-19/Denmark/DCGC-14344/2020|EPI_ISL_711707|202 ···················· ···················· ····················· ···················· ···················· ····················
X 3849 hCoV-19/Denmark/DCGC-14345/2020|EPI_ISL_711708|202 ···················· ···················· ····················· ···················· ···················· ····················
X 3850 hCoV-19/Denmark/DCGC-14347/2020|EPI_ISL_711709|202 ···················· ···················· ····················· ···················· ···················· ····················
X 3851 hCoV-19/Denmark/DCGC-14348/2020|EPI_ISL_711710|202 ···················· ···················· ····················· ···················· ···················· ····················
X 3852 hCoV-19/Denmark/DCGC-14349/2020|EPI_ISL_711711|202 ···················· ···················· ····················· ···················· ···················· ····················
X 3853 hCoV-19/Denmark/DCGC-14350/2020|EPI_ISL_711712|202 ···················· ···················· ····················· ···················· ···················· ····················
X 3854 hCoV-19/Denmark/DCGC-14385/2020|EPI_ISL_711735|202 ···················· ···················· ····················· ···················· ···················· ····················
X 3855 hCoV-19/Denmark/DCGC-14386/2020|EPI_ISL_711736|202 ···················· ···················· ····················· ···················· ···················· ····················

| | | | | | | | |
|---|---|---|---|---|---|---|---|
| X | 3856 hCoV-19/Denmark/DCGC-14387/2020\|EPI_ISL_711737\|202 | ···················· | ···················· | ····················· | ···················· | ···················· | ···················· |
| X | 3857 hCoV-19/Denmark/DCGC-14388/2020\|EPI_ISL_711738\|202 | ···················· | ···················· | ····················· | ···················· | ···················· | ···················· |
| X | 3858 hCoV-19/Denmark/DCGC-14391/2020\|EPI_ISL_711739\|202 | ···················· | ···················· | ····················· | ···················· | ···················· | ···················· |
| X | 3859 hCoV-19/Denmark/DCGC-14394/2020\|EPI_ISL_711740\|202 | ···················· | ···················· | ····················· | ···················· | ···················· | ···················· |
| X | 3860 hCoV-19/Denmark/DCGC-14395/2020\|EPI_ISL_711741\|202 | ···················· | ···················· | ····················· | ···················· | ···················· | ···················· |
| X | 3861 hCoV-19/Denmark/DCGC-14396/2020\|EPI_ISL_711742\|202 | ···················· | ···················· | ····················· | ···················· | ···················· | ···················· |
| X | 3862 hCoV-19/Denmark/DCGC-14397/2020\|EPI_ISL_711743\|202 | ···················· | ···················· | ····················· | ···················· | ···················· | ···················· |
| X | 3863 hCoV-19/Denmark/DCGC-14399/2020\|EPI_ISL_711744\|202 | ···················· | ···················· | ····················· | ···················· | ···················· | ···················· |
| X | 3864 hCoV-19/Denmark/DCGC-14401/2020\|EPI_ISL_711745\|202 | ···················· | ···················· | ····················· | ···················· | ···················· | ···················· |
| X | 3865 hCoV-19/Denmark/DCGC-14402/2020\|EPI_ISL_711746\|202 | ···················· | ···················· | ····················· | ···················· | ···················· | ···················· |
| X | 3866 hCoV-19/Denmark/DCGC-14404/2020\|EPI_ISL_711747\|202 | ···················· | ···················· | ····················· | ···················· | ···················· | ···················· |
| X | 3867 hCoV-19/Denmark/DCGC-14405/2020\|EPI_ISL_711748\|202 | ···················· | ···················· | ····················· | ···················· | ···················· | ···················· |
| X | 3868 hCoV-19/Denmark/DCGC-14407/2020\|EPI_ISL_711749\|202 | ···················· | ···················· | ····················· | ···················· | ···················· | ···················· |
| X | 3869 hCoV-19/Denmark/DCGC-14408/2020\|EPI_ISL_711750\|202 | ···················· | ···················· | ····················· | ···················· | ···················· | ···················· |
| X | 3870 hCoV-19/Denmark/DCGC-14410/2020\|EPI_ISL_711751\|202 | ···················· | ···················· | ····················· | ···················· | ···················· | ···················· |
| X | 3871 hCoV-19/Denmark/DCGC-14411/2020\|EPI_ISL_711752\|202 | ···················· | ···················· | ····················· | ···················· | ···················· | ···················· |
| X | 3872 hCoV-19/Denmark/DCGC-14412/2020\|EPI_ISL_711753\|202 | ···················· | ···················· | ····················· | ···················· | ···················· | ···················· |
| X | 3873 hCoV-19/Denmark/DCGC-14416/2020\|EPI_ISL_711754\|202 | ···················· | ···················· | ····················· | ···················· | ···················· | ···················· |
| X | 3874 hCoV-19/Denmark/DCGC-14417/2020\|EPI_ISL_711755\|202 | ···················· | ···················· | ····················· | ···················· | ···················· | ···················· |
| X | 3875 hCoV-19/Denmark/DCGC-14418/2020\|EPI_ISL_711756\|202 | ···················· | ···················· | ····················· | ···················· | ···················· | ···················· |
| X | 3876 hCoV-19/Denmark/DCGC-15618/2020\|EPI_ISL_713400\|202 | ···················· | ···················· | ····················· | ···················· | ···················· | ···················· |
| X | 3877 hCoV-19/Denmark/DCGC-15620/2020\|EPI_ISL_713401\|202 | ···················· | ···················· | ····················· | ···················· | ···················· | ···················· |
| X | 3878 hCoV-19/Denmark/DCGC-15639/2020\|EPI_ISL_713402\|202 | ···················· | ···················· | ····················· | ···················· | ···················· | ···················· |
| X | 3879 hCoV-19/Denmark/DCGC-15642/2020\|EPI_ISL_713403\|202 | ···················· | ···················· | ····················· | ···················· | ···················· | ···················· |
| X | 3880 hCoV-19/Denmark/DCGC-15675/2020\|EPI_ISL_713404\|202 | ···················· | ···················· | ····················· | ···················· | ···················· | ···················· |
| X | 3881 hCoV-19/Denmark/DCGC-15702/2020\|EPI_ISL_713405\|202 | ···················· | ···················· | ····················· | ···················· | ···················· | ···················· |
| X | 3882 hCoV-19/Denmark/DCGC-15704/2020\|EPI_ISL_713406\|202 | ···················· | ···················· | ····················· | ···················· | ···················· | ···················· |
| X | 3883 hCoV-19/Denmark/DCGC-15708/2020\|EPI_ISL_713407\|202 | ···················· | ···················· | ····················· | ···················· | ···················· | ···················· |
| X | 3884 hCoV-19/Denmark/DCGC-15721/2020\|EPI_ISL_713408\|202 | ···················· | ···················· | ····················· | ···················· | ···················· | ···················· |
| X | 3885 hCoV-19/Denmark/DCGC-15724/2020\|EPI_ISL_713409\|202 | ···················· | ···················· | ····················· | ···················· | ···················· | ···················· |
| X | 3886 hCoV-19/Denmark/DCGC-15735/2020\|EPI_ISL_713410\|202 | ···················· | ···················· | ····················· | ···················· | ···················· | ···················· |
| X | 3887 hCoV-19/Denmark/DCGC-15757/2020\|EPI_ISL_713411\|202 | ···················· | ···················· | ····················· | ···················· | ···················· | ···················· |
| X | 3888 hCoV-19/Denmark/DCGC-15760/2020\|EPI_ISL_713412\|202 | ···················· | ···················· | ····················· | ···················· | ···················· | ···················· |
| X | 3889 hCoV-19/Denmark/DCGC-15772/2020\|EPI_ISL_713413\|202 | ···················· | ···················· | ····················· | ···················· | ···················· | ···················· |
| X | 3890 hCoV-19/Denmark/DCGC-15784/2020\|EPI_ISL_713414\|202 | ···················· | ···················· | ····················· | ···················· | ···················· | ···················· |
| X | 3891 hCoV-19/Denmark/DCGC-15786/2020\|EPI_ISL_713415\|202 | ···················· | ···················· | ····················· | ···················· | ···················· | ···················· |
| X | 3892 hCoV-19/Denmark/DCGC-15831/2020\|EPI_ISL_713416\|202 | ···················· | ···················· | ····················· | ···················· | ···················· | ···················· |
| X | 3893 hCoV-19/Denmark/DCGC-15849/2020\|EPI_ISL_713417\|202 | ···················· | ···················· | ····················· | ···················· | ···················· | ···················· |
| X | 3894 hCoV-19/Denmark/DCGC-15861/2020\|EPI_ISL_713418\|202 | ···················· | ···················· | ····················· | ···················· | ···················· | ···················· |
| X | 3895 hCoV-19/Denmark/DCGC-15865/2020\|EPI_ISL_713419\|202 | ···················· | ···················· | ····················· | ···················· | ···················· | ···················· |
| X | 3896 hCoV-19/Denmark/DCGC-15877/2020\|EPI_ISL_713420\|202 | ···················· | ···················· | ····················· | ···················· | ···················· | ···················· |
| X | 3897 hCoV-19/Denmark/DCGC-15881/2020\|EPI_ISL_713421\|202 | ···················· | ···················· | ····················· | ···················· | ···················· | ···················· |
| X | 3898 hCoV-19/Denmark/DCGC-15882/2020\|EPI_ISL_713422\|202 | ···················· | ···················· | ····················· | ···················· | ···················· | ···················· |
| X | 3899 hCoV-19/Denmark/DCGC-15892/2020\|EPI_ISL_713423\|202 | ···················· | ···················· | ····················· | ···················· | ···················· | ···················· |
| X | 3900 hCoV-19/Denmark/DCGC-15319/2020\|EPI_ISL_713425\|202 | ···················· | ···················· | ····················· | ···················· | ···················· | ··A················· |
| X | 3901 hCoV-19/Denmark/DCGC-15390/2020\|EPI_ISL_713426\|202 | ···················· | ···················· | ····················· | ···················· | ···················· | ···················· |
| X | 3902 hCoV-19/Denmark/DCGC-15531/2020\|EPI_ISL_713427\|202 | ···················· | ·A·················· | ····················· | ···················· | ···················· | ··A················· |
| X | 3903 hCoV-19/Denmark/DCGC-15609/2020\|EPI_ISL_713428\|202 | ···················· | ···················· | ····················· | ···················· | ·····T·············· | ···················· |
| X | 3904 hCoV-19/Denmark/DCGC-15891/2020\|EPI_ISL_713429\|202 | ···················· | ···················· | ····················· | ···················· | ···················· | ···················· |
| X | 3905 hCoV-19/Denmark/DCGC-16318/2020\|EPI_ISL_714700\|202 | ···················· | ···················· | ····················· | ···················· | ···················· | ···················· |
| X | 3906 hCoV-19/Denmark/DCGC-16320/2020\|EPI_ISL_714701\|202 | ···················· | ···················· | ····················· | ···················· | ···················· | ···················· |
| X | 3907 hCoV-19/Denmark/DCGC-16321/2020\|EPI_ISL_714702\|202 | ···················· | ···················· | ····················· | ···················· | ···················· | ···················· |
| X | 3908 hCoV-19/Denmark/DCGC-16324/2020\|EPI_ISL_714703\|202 | ···················· | ···················· | ····················· | ···················· | ···················· | ···················· |
| X | 3909 hCoV-19/Denmark/DCGC-16328/2020\|EPI_ISL_714705\|202 | ···················· | ···················· | ····················· | ···················· | ···················· | ···················· |
| X | 3910 hCoV-19/Denmark/DCGC-16330/2020\|EPI_ISL_714706\|202 | ···················· | ···················· | ····················· | ···················· | ···················· | ···················· |
| X | 3911 hCoV-19/Denmark/DCGC-16331/2020\|EPI_ISL_714707\|202 | ···················· | ···················· | ····················· | ···················· | ···················· | ···················· |
| X | 3912 hCoV-19/Denmark/DCGC-16333/2020\|EPI_ISL_714708\|202 | ···················· | ···················· | ····················· | ···················· | ···················· | ···················· |
| X | 3913 hCoV-19/Denmark/DCGC-16335/2020\|EPI_ISL_714709\|202 | ···················· | ···················· | ····················· | ···················· | ···················· | ···················· |
| X | 3914 hCoV-19/Denmark/DCGC-16337/2020\|EPI_ISL_714710\|202 | ···················· | ···················· | ····················· | ···················· | ···················· | ···················· |
| X | 3915 hCoV-19/Denmark/DCGC-16338/2020\|EPI_ISL_714711\|202 | ···················· | ···················· | ····················· | ···················· | ···················· | ···················· |
| X | 3916 hCoV-19/Denmark/DCGC-16339/2020\|EPI_ISL_714712\|202 | ···················· | ···················· | ····················· | ···················· | ···················· | ···················· |
| X | 3917 hCoV-19/Denmark/DCGC-16340/2020\|EPI_ISL_714713\|202 | ···················· | ···················· | ····················· | ···················· | ·····T·············· | ···················· |
| X | 3918 hCoV-19/Denmark/DCGC-16341/2020\|EPI_ISL_714714\|202 | ···················· | ···················· | ····················· | ···················· | ···················· | ···················· |
| X | 3919 hCoV-19/Denmark/DCGC-16345/2020\|EPI_ISL_714715\|202 | ···················· | ···················· | ····················· | ···················· | ···················· | ···················· |
| X | 3920 hCoV-19/Denmark/DCGC-16349/2020\|EPI_ISL_714716\|202 | ···················· | ···················· | ····················· | ···················· | ···················· | ···················· |
| X | 3921 hCoV-19/Denmark/DCGC-16351/2020\|EPI_ISL_714717\|202 | ···················· | ···················· | ····················· | ···················· | ···················· | ···················· |
| X | 3922 hCoV-19/Denmark/DCGC-16353/2020\|EPI_ISL_714718\|202 | ···················· | ···················· | ····················· | ···················· | ···················· | ···················· |
| X | 3923 hCoV-19/Denmark/DCGC-16354/2020\|EPI_ISL_714719\|202 | ···················· | ···················· | ····················· | ···················· | ···················· | ···················· |
| X | 3924 hCoV-19/Denmark/DCGC-16357/2020\|EPI_ISL_714720\|202 | ···················· | ···················· | ····················· | ···················· | ···················· | ···················· |
| X | 3925 hCoV-19/Denmark/DCGC-16358/2020\|EPI_ISL_714721\|202 | ···················· | ···················· | ····················· | ···················· | ···················· | ···················· |
| X | 3926 hCoV-19/Denmark/DCGC-16360/2020\|EPI_ISL_714722\|202 | ···················· | ···················· | ····················· | ···················· | ···················· | ···················· |
| X | 3927 hCoV-19/Denmark/DCGC-16362/2020\|EPI_ISL_714723\|202 | ···················· | ···················· | ····················· | ···················· | ···················· | ···················· |
| X | 3928 hCoV-19/Denmark/DCGC-16363/2020\|EPI_ISL_714724\|202 | ···················· | ···················· | ····················· | ···················· | ···················· | ···················· |
| X | 3929 hCoV-19/Denmark/DCGC-16364/2020\|EPI_ISL_714725\|202 | ···················· | ···················· | ····················· | ···················· | ···················· | ···················· |
| X | 3930 hCoV-19/Denmark/DCGC-16366/2020\|EPI_ISL_714726\|202 | ···················· | ···················· | ····················· | ···················· | ···················· | ···················· |
| X | 3931 hCoV-19/Denmark/DCGC-16367/2020\|EPI_ISL_714727\|202 | ···················· | ···················· | ····················· | ···················· | ···················· | ···················· |
| X | 3932 hCoV-19/Denmark/DCGC-16369/2020\|EPI_ISL_714728\|202 | ···················· | ···················· | ····················· | ···················· | ···················· | ···················· |
| X | 3933 hCoV-19/Denmark/DCGC-16370/2020\|EPI_ISL_714729\|202 | ···················· | ·A·················· | ····················· | ···················· | ···················· | ···················· |
| X | 3934 hCoV-19/Denmark/DCGC-16372/2020\|EPI_ISL_714730\|202 | ···················· | ···················· | ····················· | ···················· | ···················· | ···················· |
| X | 3935 hCoV-19/Denmark/DCGC-16373/2020\|EPI_ISL_714731\|202 | ···················· | ···················· | ····················· | ···················· | ···················· | ···················· |

| | | | | | | | |
|---|---|---|---|---|---|---|---|
| X | 3936 hCoV-19/Denmark/DCGC-16375/2020\|EPI_ISL_714732\|202 | ···················· | ···················· | ····················· | ···················· | ···················· | ···················· |
| X | 3937 hCoV-19/Denmark/DCGC-16377/2020\|EPI_ISL_714733\|202 | ···················· | ···················· | ····················· | ···················· | ···················· | ···················· |
| X | 3938 hCoV-19/Denmark/DCGC-16378/2020\|EPI_ISL_714734\|202 | ···················· | ···················· | ····················· | ···················· | ···················· | ···················· |
| X | 3939 hCoV-19/Denmark/DCGC-16383/2020\|EPI_ISL_714735\|202 | ···················· | ···················· | ····················· | ···················· | ···················· | ···················· |
| X | 3940 hCoV-19/Denmark/DCGC-16386/2020\|EPI_ISL_714736\|202 | ···················· | ···················· | ····················· | ···················· | ···················· | ···················· |
| X | 3941 hCoV-19/Denmark/DCGC-16387/2020\|EPI_ISL_714737\|202 | ···················· | ···················· | ····················· | ···················· | ···················· | ···················· |
| X | 3942 hCoV-19/Denmark/DCGC-16392/2020\|EPI_ISL_714738\|202 | ···················· | ···················· | ····················· | ···················· | ···················· | ···················· |
| X | 3943 hCoV-19/Denmark/DCGC-16393/2020\|EPI_ISL_714739\|202 | ···················· | ···················· | ····················· | ···················· | ···················· | ···················· |
| X | 3944 hCoV-19/Denmark/DCGC-16394/2020\|EPI_ISL_714740\|202 | ···················· | ···················· | ····················· | ···················· | ···················· | ···················· |
| X | 3945 hCoV-19/Denmark/DCGC-16395/2020\|EPI_ISL_714741\|202 | ···················· | ···················· | ····················· | ···················· | ······T············· | ···················· |
| X | 3946 hCoV-19/Denmark/DCGC-16396/2020\|EPI_ISL_714742\|202 | ···················· | ···················· | ····················· | ···················· | ···················· | ···················· |
| X | 3947 hCoV-19/Denmark/DCGC-16398/2020\|EPI_ISL_714743\|202 | ···················· | ···················· | ····················· | ···················· | ···················· | ···················· |
| X | 3948 hCoV-19/Denmark/DCGC-16401/2020\|EPI_ISL_714744\|202 | ···················· | ···················· | ····················· | ···················· | ···················· | ···················· |
| X | 3949 hCoV-19/Denmark/DCGC-16402/2020\|EPI_ISL_714745\|202 | ···················· | ···················· | ····················· | ···················· | ···················· | ···················· |
| X | 3950 hCoV-19/Denmark/DCGC-16404/2020\|EPI_ISL_714746\|202 | ···················· | ·A·················· | ····················· | ···················· | ···················· | ···A················ |
| X | 3951 hCoV-19/Denmark/DCGC-16405/2020\|EPI_ISL_714747\|202 | ···················· | ···················· | ····················· | ···················· | ···················· | ···················· |
| X | 3952 hCoV-19/Denmark/DCGC-16406/2020\|EPI_ISL_714748\|202 | ···················· | ···················· | ····················· | ···················· | ···················· | ···················· |
| X | 3953 hCoV-19/Denmark/DCGC-16409/2020\|EPI_ISL_714749\|202 | ···················· | ···················· | ····················· | ···················· | ···················· | ···················· |
| X | 3954 hCoV-19/Denmark/DCGC-16410/2020\|EPI_ISL_714750\|202 | ···················· | ···················· | ····················· | ···················· | ···················· | ···················· |
| X | 3955 hCoV-19/Denmark/DCGC-16411/2020\|EPI_ISL_714751\|202 | ···················· | ···················· | ····················· | ···················· | ···················· | ···················· |
| X | 3956 hCoV-19/Denmark/DCGC-16413/2020\|EPI_ISL_714752\|202 | ···················· | ···················· | ····················· | ···················· | ···················· | ···················· |
| X | 3957 hCoV-19/Denmark/DCGC-16414/2020\|EPI_ISL_714753\|202 | ···················· | ···················· | ····················· | ···················· | ···················· | ···················· |
| X | 3958 hCoV-19/Denmark/DCGC-16416/2020\|EPI_ISL_714754\|202 | ···················· | ···················· | ····················· | ···················· | ···················· | ···················· |
| X | 3959 hCoV-19/Denmark/DCGC-16419/2020\|EPI_ISL_714755\|202 | ···················· | ···················· | ····················· | ···················· | ···················· | ···················· |
| X | 3960 hCoV-19/Denmark/DCGC-16421/2020\|EPI_ISL_714756\|202 | ···················· | ···················· | ····················· | ···················· | ···················· | ···················· |
| X | 3961 hCoV-19/Denmark/DCGC-16423/2020\|EPI_ISL_714757\|202 | ···················· | ···················· | ····················· | ···················· | ···················· | ···················· |
| X | 3962 hCoV-19/Denmark/DCGC-16425/2020\|EPI_ISL_714758\|202 | ···················· | ···················· | ····················· | ···················· | ···················· | ···················· |
| X | 3963 hCoV-19/Denmark/DCGC-16426/2020\|EPI_ISL_714759\|202 | ···················· | ···················· | ····················· | ···················· | ···················· | ···················· |
| X | 3964 hCoV-19/Denmark/DCGC-16429/2020\|EPI_ISL_714760\|202 | ···················· | ···················· | ····················· | ···················· | ···················· | ···················· |
| X | 3965 hCoV-19/Denmark/DCGC-16430/2020\|EPI_ISL_714761\|202 | ···················· | ···················· | ····················· | ···················· | ···················· | ···················· |
| X | 3966 hCoV-19/Denmark/DCGC-16431/2020\|EPI_ISL_714762\|202 | ···················· | ···················· | ····················· | ···················· | ···················· | ···················· |
| X | 3967 hCoV-19/Denmark/DCGC-16432/2020\|EPI_ISL_714763\|202 | ···················· | ···················· | ····················· | ···················· | ···················· | ···················· |
| X | 3968 hCoV-19/Denmark/DCGC-16435/2020\|EPI_ISL_714764\|202 | ···················· | ···················· | ····················· | ···················· | ···················· | ···················· |
| X | 3969 hCoV-19/Denmark/DCGC-16436/2020\|EPI_ISL_714765\|202 | ···················· | ···················· | ····················· | ···················· | ···················· | ···················· |
| X | 3970 hCoV-19/Denmark/DCGC-16437/2020\|EPI_ISL_714766\|202 | ···················· | ···················· | ····················· | ···················· | ···················· | ···················· |
| X | 3971 hCoV-19/Denmark/DCGC-16438/2020\|EPI_ISL_714767\|202 | ···················· | ···················· | ····················· | ···················· | ···················· | ···················· |
| X | 3972 hCoV-19/Denmark/DCGC-16439/2020\|EPI_ISL_714768\|202 | ···················· | ···················· | ····················· | ···················· | ···················· | ···················· |
| X | 3973 hCoV-19/Denmark/DCGC-16440/2020\|EPI_ISL_714769\|202 | ···················· | ···················· | ····················· | ···················· | ···················· | ···················· |
| X | 3974 hCoV-19/Denmark/DCGC-16441/2020\|EPI_ISL_714770\|202 | ···················· | ···················· | ····················· | ···················· | ···················· | ···················· |
| X | 3975 hCoV-19/Denmark/DCGC-16443/2020\|EPI_ISL_714771\|202 | ···················· | ···················· | ····················· | ···················· | ···················· | ···················· |
| X | 3976 hCoV-19/Denmark/DCGC-16446/2020\|EPI_ISL_714772\|202 | ···················· | ···················· | ····················· | ···················· | ···················· | ···················· |
| X | 3977 hCoV-19/Denmark/DCGC-16449/2020\|EPI_ISL_714773\|202 | ···················· | ···················· | ····················· | ···················· | ···················· | ···················· |
| X | 3978 hCoV-19/Denmark/DCGC-16450/2020\|EPI_ISL_714774\|202 | ···················· | ···················· | ····················· | ···················· | ···················· | ···················· |
| X | 3979 hCoV-19/Denmark/DCGC-16452/2020\|EPI_ISL_714775\|202 | ···················· | ·A·················· | ····················· | ···················· | ···················· | ···················· |
| X | 3980 hCoV-19/Denmark/DCGC-16453/2020\|EPI_ISL_714776\|202 | ···················· | ···················· | ····················· | ···················· | ···················· | ···················· |
| X | 3981 hCoV-19/Denmark/DCGC-16454/2020\|EPI_ISL_714777\|202 | ···················· | ···················· | ····················· | ···················· | ···················· | ···················· |
| X | 3982 hCoV-19/Denmark/DCGC-16456/2020\|EPI_ISL_714778\|202 | ···················· | ···················· | ····················· | ···················· | ···················· | ···················· |
| X | 3983 hCoV-19/Denmark/DCGC-16458/2020\|EPI_ISL_714779\|202 | ···················· | ···················· | ····················· | ···················· | ···················· | ···················· |
| X | 3984 hCoV-19/Denmark/DCGC-16460/2020\|EPI_ISL_714780\|202 | ···················· | ···················· | ····················· | ···················· | ···················· | ···················· |
| X | 3985 hCoV-19/Denmark/DCGC-16463/2020\|EPI_ISL_714781\|202 | ···················· | ···················· | ····················· | ···················· | ···················· | ···················· |
| X | 3986 hCoV-19/Denmark/DCGC-16465/2020\|EPI_ISL_714782\|202 | ···················· | ···················· | ····················· | ···················· | ···················· | ···················· |
| X | 3987 hCoV-19/Denmark/DCGC-16466/2020\|EPI_ISL_714783\|202 | ···················· | ···················· | ····················· | ···················· | ···················· | ···················· |
| X | 3988 hCoV-19/Denmark/DCGC-16467/2020\|EPI_ISL_714784\|202 | ···················· | ·A·················· | ····················· | ···················· | ···················· | ···················· |
| X | 3989 hCoV-19/Denmark/DCGC-16468/2020\|EPI_ISL_714785\|202 | ···················· | ···················· | ····················· | ···················· | ···················· | ···················· |
| X | 3990 hCoV-19/Denmark/DCGC-16469/2020\|EPI_ISL_714786\|202 | ···················· | ···················· | ····················· | ···················· | ···················· | ···················· |
| X | 3991 hCoV-19/Denmark/DCGC-16470/2020\|EPI_ISL_714787\|202 | ···················· | ···················· | ····················· | ···················· | ···················· | ···················· |
| X | 3992 hCoV-19/Denmark/DCGC-16471/2020\|EPI_ISL_714788\|202 | ···················· | ···················· | ····················· | ···················· | ···················· | ···················· |
| X | 3993 hCoV-19/Denmark/DCGC-16473/2020\|EPI_ISL_714789\|202 | ···················· | ···················· | ····················· | ···················· | ···················· | ···················· |
| X | 3994 hCoV-19/Denmark/DCGC-16474/2020\|EPI_ISL_714790\|202 | ···················· | ···················· | ····················· | ···················· | ···················· | ···················· |
| X | 3995 hCoV-19/Denmark/DCGC-16476/2020\|EPI_ISL_714791\|202 | ···················· | ···················· | ····················· | ···················· | ···················· | ···················· |
| X | 3996 hCoV-19/Denmark/DCGC-16478/2020\|EPI_ISL_714792\|202 | ···················· | ···················· | ····················· | ···················· | ···················· | ···················· |
| X | 3997 hCoV-19/Denmark/DCGC-16481/2020\|EPI_ISL_714793\|202 | ···················· | ·A·················· | ····················· | ···················· | ···················· | ···················· |
| X | 3998 hCoV-19/Denmark/DCGC-16483/2020\|EPI_ISL_714794\|202 | ···················· | ···················· | ····················· | ···················· | ···················· | ···················· |
| X | 3999 hCoV-19/Denmark/DCGC-16484/2020\|EPI_ISL_714795\|202 | ···················· | ···················· | ····················· | ···················· | ···················· | ···················· |
| X | 4000 hCoV-19/Denmark/DCGC-16487/2020\|EPI_ISL_714796\|202 | ···················· | ···················· | ····················· | ···················· | ···················· | ···················· |
| X | 4001 hCoV-19/Denmark/DCGC-16488/2020\|EPI_ISL_714797\|202 | ···················· | ···················· | ····················· | ···················· | ···················· | ···················· |
| X | 4002 hCoV-19/Denmark/DCGC-16489/2020\|EPI_ISL_714798\|202 | ···················· | ···················· | ····················· | ···················· | ···················· | ···················· |
| X | 4003 hCoV-19/Denmark/DCGC-16490/2020\|EPI_ISL_714799\|202 | ···················· | ···················· | ····················· | ···················· | ···················· | ···················· |
| X | 4004 hCoV-19/Malaysia/IMR-WI115/2020\|EPI_ISL_718272\|202 | ···················· | ···················· | ····················· | ···················· | ···················· | ···················· |
| X | 4005 hCoV-19/Japan/FHU-CS24-0221/2020\|EPI_ISL_718273\|20 | ···················· | ···················· | ····················· | ···················· | ···················· | ···················· |
| X | 4006 hCoV-19/Malaysia/IMR-WI111/2020\|EPI_ISL_718274\|202 | ···················· | ···················· | ····················· | ···················· | ···················· | ···················· |
| X | 4007 hCoV-19/Malaysia/IMR-WI119/2020\|EPI_ISL_718275\|202 | ···················· | ···················· | ····················· | ···················· | ···················· | ···················· |
| X | 4008 hCoV-19/Malaysia/IMR-WI120/2020\|EPI_ISL_718276\|202 | ···················· | ···················· | ····················· | ···················· | ···················· | ···················· |
| X | 4009 hCoV-19/Malaysia/IMR-WI123/2020\|EPI_ISL_718277\|202 | ···················· | ···················· | ····················· | ···················· | ···················· | ···················· |
| X | 4010 hCoV-19/Malaysia/IMR-WI132/2020\|EPI_ISL_718280\|202 | ···················· | ···················· | ····················· | ···················· | ···················· | ···················· |
| X | 4011 hCoV-19/Malaysia/IMR-WI133/2020\|EPI_ISL_718281\|202 | ···················· | ···················· | ····················· | ···················· | ···················· | ···················· |
| X | 4012 hCoV-19/Malaysia/IMR-WI147/2020\|EPI_ISL_718282\|202 | ···················· | ···················· | ····················· | ···················· | ···················· | ···················· |
| X | 4013 hCoV-19/Malaysia/IMR-WI109/2020\|EPI_ISL_718283\|202 | ···················· | ···················· | ····················· | ···················· | ···················· | ···················· |
| X | 4014 hCoV-19/Malaysia/IMR-WC124489/2020\|EPI_ISL_718284\| | ···················· | ···················· | ····················· | ···················· | ···················· | ···················· |
| X | 4015 hCoV-19/Malaysia/IMR-WI203/2020\|EPI_ISL_718285\|202 | ···················· | ···················· | ····················· | ···················· | ···················· | ···················· |

```
X  4016 hCoV-19/Malaysia/IMR-CV136859/2020|EPI_ISL_718287|  ····················  ·A··················  ·····················  ····················  ····················  ····················
X  4017 hCoV-19/England/MILK-BABF5A/2020|EPI_ISL_720590|20  ····················  ······A·············  ·····················  ····················  ····················  ····················
X  4018 hCoV-19/England/MILK-BAC566/2020|EPI_ISL_720591|20  ····················  ····················  ·····················  ····················  ····················  ····················
X  4019 hCoV-19/England/MILK-BB1AE7/2020|EPI_ISL_720592|20  ····················  ····················  ·····················  ····················  ····················  ····················
X  4020 hCoV-19/England/MILK-BB081B/2020|EPI_ISL_720593|20  ····················  ····················  ·····················  ····················  ····················  ····················
X  4021 hCoV-19/England/MILK-BB10C7/2020|EPI_ISL_720594|20  ····················  ····················  ·····················  ····················  ····················  ····················
X  4022 hCoV-19/England/MILK-BB19BD/2020|EPI_ISL_720595|20  ····················  ····················  ·····················  ····················  ····················  ····················
X  4023 hCoV-19/England/MILK-BB1DEE/2020|EPI_ISL_720596|20  ····················  ····················  ·····················  ····················  ····················  ····················
X  4024 hCoV-19/England/MILK-BB1B99/2020|EPI_ISL_720598|20  ····················  ····················  ·····················  ····················  ····················  ····················
X  4025 hCoV-19/England/MILK-BB22C0/2020|EPI_ISL_720599|20  ····················  ····················  ·····················  ····················  ····················  ····················
X  4026 hCoV-19/Switzerland/SZ-ETHZ-340144/2020|EPI_ISL_72  ····················  ····················  ·····················  ····················  ····················  ····················
X  4027 hCoV-19/Switzerland/SZ-ETHZ-390388/2020|EPI_ISL_72  ····················  ····················  ·····················  ····················  ····················  ····················
X  4028 hCoV-19/Switzerland/SZ-ETHZ-390396/2020|EPI_ISL_72  ····················  ····················  ·····················  ····················  ······K·············  ····················
X  4029 hCoV-19/Switzerland/SZ-ETHZ-390407/2020|EPI_ISL_72  ····················  ····················  ·····················  ····················  ····················  ····················
X  4030 hCoV-19/Switzerland/SZ-ETHZ-390410/2020|EPI_ISL_72  ····················  ····················  ·····················  ····················  ····················  ····················
X  4031 hCoV-19/Switzerland/TG-ETHZ-380304/2020|EPI_ISL_72  ····················  ······A·············  ·····················  ····················  ····················  ····················
X  4032 hCoV-19/Switzerland/TG-ETHZ-390379/2020|EPI_ISL_72  ········Y···········  ····················  ·····················  ····················  ····················  ····················
X  4033 hCoV-19/Switzerland/TG-ETHZ-390380/2020|EPI_ISL_72  ····················  ····················  ·····················  ····················  ····················  ····················
X  4034 hCoV-19/Switzerland/TG-ETHZ-390401/2020|EPI_ISL_72  ····················  ····················  ·····················  ····················  ····················  ····················
X  4035 hCoV-19/Switzerland/TG-ETHZ-390404/2020|EPI_ISL_72  ····················  ··G·················  ·····················  ····················  ····················  ····················
X  4036 hCoV-19/Wales/ALDP-B75E89/2020|EPI_ISL_723714|2020  ····················  ····················  ·····················  ····················  ····················  ····················
X  4037 hCoV-19/England/ALDP-B75EE3/2020|EPI_ISL_723715|20  ····················  ····················  ·····················  ····················  ····················  ····················
X  4038 hCoV-19/England/ALDP-B75F2C/2020|EPI_ISL_723716|20  ····················  ····················  ·····················  ····················  ····················  ····················
X  4039 hCoV-19/England/ALDP-B75F77/2020|EPI_ISL_723717|20  ····················  ····················  ·····················  ····················  ····················  ····················
X  4040 hCoV-19/England/ALDP-B75F95/2020|EPI_ISL_723718|20  ····················  ····················  ·····················  ····················  ····················  ····················
X  4041 hCoV-19/England/ALDP-B75FB3/2020|EPI_ISL_723719|20  ····················  ····················  ·····················  ····················  ····················  ····················
X  4042 hCoV-19/England/ALDP-B75FC2/2020|EPI_ISL_723720|20  ····················  ····················  ·····················  ····················  ····················  ····················
X  4043 hCoV-19/England/ALDP-B75FD1/2020|EPI_ISL_723721|20  ····················  ····················  ·····················  ····················  ····················  ····················
X  4044 hCoV-19/England/ALDP-B75FE0/2020|EPI_ISL_723722|20  ····················  ····················  ·····················  ····················  ····················  ····················
X  4045 hCoV-19/England/ALDP-B75FFF/2020|EPI_ISL_723723|20  ····················  ····················  ·····················  ····················  ····················  ····················
X  4046 hCoV-19/England/ALDP-B76055/2020|EPI_ISL_723726|20  ····················  ····················  ·····················  ····················  ····················  ····················
X  4047 hCoV-19/England/ALDP-B76082/2020|EPI_ISL_723729|20  ····················  ····················  ·····················  ····················  ····················  ····················
X  4048 hCoV-19/England/ALDP-B76091/2020|EPI_ISL_723730|20  ····················  ····················  ·····················  ····················  ····················  ····················
X  4049 hCoV-19/England/ALDP-B760DD/2020|EPI_ISL_723731|20  ····················  ····················  ·····················  ····················  ····················  ····················
X  4050 hCoV-19/England/ALDP-B760EC/2020|EPI_ISL_723732|20  ····················  ····················  ·····················  ····················  ····················  ····················
X  4051 hCoV-19/England/ALDP-B760FB/2020|EPI_ISL_723733|20  ····················  ····················  ·····················  ····················  ····················  ····················
X  4052 hCoV-19/England/ALDP-B76107/2020|EPI_ISL_723734|20  ····················  ····················  ·····················  ····················  ····················  ····················
X  4053 hCoV-19/England/ALDP-B76116/2020|EPI_ISL_723735|20  ····················  ····················  ·····················  ····················  ····················  ····················
X  4054 hCoV-19/England/CAMC-BBBC23/2020|EPI_ISL_724100|20  ····················  ····················  ·····················  ····················  ····················  ····················
X  4055 hCoV-19/England/CAMC-BBBC41/2020|EPI_ISL_724102|20  ····················  ····················  ·····················  ····················  ····················  ····················
X  4056 hCoV-19/England/CAMC-BBBC50/2020|EPI_ISL_724103|20  ····················  ····················  ·····················  ····················  ····················  ····················
X  4057 hCoV-19/England/CAMC-BBBD7B/2020|EPI_ISL_724104|20  ····················  ····················  ·····················  ····················  ····················  ····················
X  4058 hCoV-19/England/CAMC-BBBD8A/2020|EPI_ISL_724105|20  ····················  ····················  ·····················  ····················  ····················  ····················
X  4059 hCoV-19/England/CAMC-BBBDB7/2020|EPI_ISL_724107|20  ····················  ·A·······A··········  ·····················  ····················  ····················  ····················
X  4060 hCoV-19/England/CAMC-BBBDC6/2020|EPI_ISL_724108|20  ····················  ····················  ·····················  ····················  ····················  ····················
X  4061 hCoV-19/England/CAMC-BBBFC0/2020|EPI_ISL_724112|20  ····················  ····················  ·····················  ····················  ····················  ····················
X  4062 hCoV-19/England/CAMC-BBC026/2020|EPI_ISL_724118|20  ····················  ····················  ·····················  ····················  ····················  ····················
X  4063 hCoV-19/England/CAMC-BBC035/2020|EPI_ISL_724119|20  ····················  ·A··················  ·····················  ····················  ····················  ···A················
X  4064 hCoV-19/England/CAMC-BBC044/2020|EPI_ISL_724120|20  ····················  ····················  ·····················  ····················  ····················  ····················
X  4065 hCoV-19/England/CAMC-BBC071/2020|EPI_ISL_724123|20  ····················  ····················  ·····················  ····················  ····················  ····················
X  4066 hCoV-19/England/CAMC-BBC34B/2020|EPI_ISL_724125|20  ····················  ····················  ·····················  ····················  ····················  ····················
X  4067 hCoV-19/England/CAMC-BBC378/2020|EPI_ISL_724127|20  ····················  ····················  ·····················  ····················  ····················  ····················
X  4068 hCoV-19/England/CAMC-BBC387/2020|EPI_ISL_724128|20  ····················  ····················  ·····················  ····················  ····················  ····················
X  4069 hCoV-19/England/CAMC-BBC396/2020|EPI_ISL_724129|20  ····················  ····················  ·····················  ····················  ····················  ····················
X  4070 hCoV-19/England/CAMC-BBC3A5/2020|EPI_ISL_724130|20  ····················  ····················  ·····················  ····················  ····················  ····················
X  4071 hCoV-19/England/CAMC-BBC3B4/2020|EPI_ISL_724131|20  ····················  ····················  ·····················  ····················  ····················  ····················
X  4072 hCoV-19/England/CAMC-BBC3D2/2020|EPI_ISL_724133|20  ····················  ····················  ·····················  ····················  ····················  ····················
X  4073 hCoV-19/England/CAMC-BBC3F0/2020|EPI_ISL_724135|20  ····················  ·A··················  ·····················  ····················  ····················  ····················
X  4074 hCoV-19/England/CAMC-BBC439/2020|EPI_ISL_724137|20  ····················  ····················  ·····················  ····················  ····················  ····················
X  4075 hCoV-19/England/CAMC-BBC448/2020|EPI_ISL_724138|20  ····················  ····················  ·····················  ····················  ······T·············  ····················
X  4076 hCoV-19/England/CAMC-BBC493/2020|EPI_ISL_724142|20  ····················  ····················  ·····················  ····················  ····················  ····················
X  4077 hCoV-19/England/CAMC-BBC4B1/2020|EPI_ISL_724144|20  ····················  ····················  ·····················  ····················  ····················  ····················
X  4078 hCoV-19/England/CAMC-BBC4FD/2020|EPI_ISL_724148|20  ····················  ····················  ·····················  ····················  ····················  ····················
X  4079 hCoV-19/England/CAMC-BBC518/2020|EPI_ISL_724149|20  ····················  ····················  ·····················  ····················  ····················  ····················
X  4080 hCoV-19/England/CAMC-BBC527/2020|EPI_ISL_724150|20  ····················  ····················  ·····················  ····················  ····················  ····················
X  4081 hCoV-19/England/CAMC-BBC545/2020|EPI_ISL_724152|20  ····················  ····················  ·····················  ····················  ······T·············  ····················
X  4082 hCoV-19/England/CAMC-BBC581/2020|EPI_ISL_724154|20  ····················  ····················  ·····················  ····················  ····················  ····················
X  4083 hCoV-19/England/CAMC-BBC590/2020|EPI_ISL_724155|20  ····················  ····················  ·····················  ····················  ····················  ····················
X  4084 hCoV-19/England/CAMC-BBC5CD/2020|EPI_ISL_724157|20  ····················  ····················  ·····················  ····················  ····················  ····················
X  4085 hCoV-19/England/CAMC-BBC5EB/2020|EPI_ISL_724159|20  ····················  ····················  ·····················  ····················  ····················  ····················
X  4086 hCoV-19/England/CAMC-BBC606/2020|EPI_ISL_724161|20  ····················  ·A··················  ·····················  ····················  ····················  ····················
X  4087 hCoV-19/England/CAMC-BBC615/2020|EPI_ISL_724162|20  ····················  ····················  ·····················  ····················  ····················  ····················
X  4088 hCoV-19/England/CAMC-BBC67F/2020|EPI_ISL_724168|20  ····················  ····················  ·····················  ····················  ····················  ····················
X  4089 hCoV-19/England/CAMC-BBC6CA/2020|EPI_ISL_724172|20  ····················  ·A··················  ·····················  ····················  ····················  ···A················
X  4090 hCoV-19/England/CAMC-BBC6E8/2020|EPI_ISL_724174|20  ····················  ····················  ·····················  ····················  ····················  ····················
X  4091 hCoV-19/England/CAMC-BBC721/2020|EPI_ISL_724178|20  ····················  ····················  ·····················  ····················  ····················  ····················
X  4092 hCoV-19/England/CAMC-BBC730/2020|EPI_ISL_724179|20  ····················  ····················  ·····················  ····················  ····················  ····················
X  4093 hCoV-19/England/CAMC-BBC79A/2020|EPI_ISL_724184|20  ····················  ····················  ·····················  ····················  ····················  ····················
X  4094 hCoV-19/England/CAMC-BBC7A9/2020|EPI_ISL_724185|20  ····················  ····················  ·····················  ····················  ····················  ····················
X  4095 hCoV-19/England/CAMC-BBC7C7/2020|EPI_ISL_724187|20  ····················  ····················  ·····················  ····················  ····················  ····················
```

| | | | | | | | | |
|---|---|---|---|---|---|---|---|---|
| X | 4096 | hCoV-19/England/CAMC-BBC92B/2020\|EPI_ISL_724188\|20 | ···················· | ···················· | ····················· | ···················· | ···················· | ···················· |
| X | 4097 | hCoV-19/Denmark/DCGC-14351/2020\|EPI_ISL_711713\|202 | ···················· | ···················· | ····················· | ···················· | ···················· | ···················· |
| X | 4098 | hCoV-19/Denmark/DCGC-14352/2020\|EPI_ISL_711714\|202 | ···················· | ···················· | ····················· | ···················· | ···················· | ···················· |
| X | 4099 | hCoV-19/Denmark/DCGC-14354/2020\|EPI_ISL_711715\|202 | ···················· | ···················· | ····················· | ···················· | ···················· | ···················· |
| X | 4100 | hCoV-19/Denmark/DCGC-14357/2020\|EPI_ISL_711716\|202 | ···················· | ···················· | ····················· | ···················· | ···················· | ···················· |
| X | 4101 | hCoV-19/Denmark/DCGC-14358/2020\|EPI_ISL_711717\|202 | ···················· | ···················· | ····················· | ···················· | ···················· | ···················· |
| X | 4102 | hCoV-19/Denmark/DCGC-14359/2020\|EPI_ISL_711718\|202 | ···················· | ···················· | ····················· | ···················· | ···················· | ···················· |
| X | 4103 | hCoV-19/Denmark/DCGC-14361/2020\|EPI_ISL_711719\|202 | ···················· | ···················· | ····················· | ···················· | ···················· | ···················· |
| X | 4104 | hCoV-19/Denmark/DCGC-14363/2020\|EPI_ISL_711720\|202 | ···················· | ···················· | ····················· | ···················· | ···················· | ···················· |
| X | 4105 | hCoV-19/Denmark/DCGC-14364/2020\|EPI_ISL_711721\|202 | ···················· | ···················· | ····················· | ···················· | ···················· | ···················· |
| X | 4106 | hCoV-19/Denmark/DCGC-14365/2020\|EPI_ISL_711722\|202 | ···················· | ···················· | ····················· | ···················· | ···················· | ···················· |
| X | 4107 | hCoV-19/Denmark/DCGC-14366/2020\|EPI_ISL_711723\|202 | ···················· | ···················· | ····················· | ···················· | ···················· | ···················· |
| X | 4108 | hCoV-19/Denmark/DCGC-14367/2020\|EPI_ISL_711724\|202 | ···················· | ···················· | ····················· | ···················· | ···················· | ···················· |
| X | 4109 | hCoV-19/Denmark/DCGC-14368/2020\|EPI_ISL_711725\|202 | ···················· | ···················· | ····················· | ···················· | ······T············· | ···················· |
| X | 4110 | hCoV-19/Denmark/DCGC-14369/2020\|EPI_ISL_711726\|202 | ···················· | ···················· | ····················· | ···················· | ···················· | ···················· |
| X | 4111 | hCoV-19/Denmark/DCGC-14371/2020\|EPI_ISL_711727\|202 | ···················· | ···················· | ····················· | ···················· | ···················· | ···················· |
| X | 4112 | hCoV-19/Denmark/DCGC-14372/2020\|EPI_ISL_711728\|202 | ···················· | ···················· | ····················· | ···················· | ···················· | ···················· |
| X | 4113 | hCoV-19/Denmark/DCGC-14375/2020\|EPI_ISL_711729\|202 | ···················· | ···················· | ····················· | ···················· | ···················· | ···················· |
| X | 4114 | hCoV-19/Denmark/DCGC-14378/2020\|EPI_ISL_711730\|202 | ···················· | ···················· | ····················· | ···················· | ···················· | ···················· |
| X | 4115 | hCoV-19/Denmark/DCGC-14380/2020\|EPI_ISL_711731\|202 | ···················· | ···················· | ····················· | ···················· | ···················· | ···················· |
| X | 4116 | hCoV-19/Denmark/DCGC-14381/2020\|EPI_ISL_711732\|202 | ···················· | ···················· | ····················· | ···················· | ···················· | ···················· |
| X | 4117 | hCoV-19/Denmark/DCGC-14383/2020\|EPI_ISL_711733\|202 | ···················· | ·A·················· | ····················· | ···················· | ···················· | ···················· |
| X | 4118 | hCoV-19/Denmark/DCGC-14384/2020\|EPI_ISL_711734\|202 | ···················· | ···················· | ····················· | ···················· | ···················· | ···················· |
| X | 4119 | hCoV-19/Denmark/DCGC-14421/2020\|EPI_ISL_711757\|202 | ···················· | ···················· | ····················· | ···················· | ···················· | ···················· |
| X | 4120 | hCoV-19/Denmark/DCGC-14424/2020\|EPI_ISL_711758\|202 | ···················· | ···················· | ····················· | ···················· | ···················· | ···················· |
| X | 4121 | hCoV-19/Denmark/DCGC-14426/2020\|EPI_ISL_711759\|202 | ···················· | ···················· | ····················· | ···················· | ···················· | ···················· |
| X | 4122 | hCoV-19/Denmark/DCGC-14427/2020\|EPI_ISL_711760\|202 | ···················· | ···················· | ····················· | ···················· | ···················· | ···················· |
| X | 4123 | hCoV-19/Denmark/DCGC-14428/2020\|EPI_ISL_711761\|202 | ···················· | ···················· | ····················· | ···················· | ···················· | ···················· |
| X | 4124 | hCoV-19/Denmark/DCGC-14430/2020\|EPI_ISL_711762\|202 | ···················· | ···················· | ····················· | ···················· | ···················· | ···················· |
| X | 4125 | hCoV-19/Denmark/DCGC-14433/2020\|EPI_ISL_711763\|202 | ···················· | ···················· | ····················· | ···················· | ···················· | ···················· |
| X | 4126 | hCoV-19/Denmark/DCGC-14434/2020\|EPI_ISL_711764\|202 | ···················· | ···················· | ····················· | ···················· | ···················· | ···················· |
| X | 4127 | hCoV-19/Denmark/DCGC-14435/2020\|EPI_ISL_711765\|202 | ···················· | ···················· | ····················· | ···················· | ···················· | ···················· |
| X | 4128 | hCoV-19/Denmark/DCGC-14436/2020\|EPI_ISL_711766\|202 | ···················· | ···················· | ····················· | ···················· | ···················· | ···················· |
| X | 4129 | hCoV-19/Denmark/DCGC-14437/2020\|EPI_ISL_711767\|202 | ···················· | ···················· | ····················· | ···················· | ···················· | ···················· |
| X | 4130 | hCoV-19/Denmark/DCGC-14438/2020\|EPI_ISL_711768\|202 | ···················· | ···················· | ····················· | ···················· | ···················· | ···················· |
| X | 4131 | hCoV-19/Denmark/DCGC-14439/2020\|EPI_ISL_711769\|202 | ···················· | ···················· | ····················· | ···················· | ···················· | ···················· |
| X | 4132 | hCoV-19/Denmark/DCGC-14440/2020\|EPI_ISL_711770\|202 | ···················· | ···················· | ····················· | ···················· | ···················· | ···················· |
| X | 4133 | hCoV-19/Denmark/DCGC-14443/2020\|EPI_ISL_711771\|202 | ···················· | ···················· | ····················· | ···················· | ···················· | ···················· |
| X | 4134 | hCoV-19/Denmark/DCGC-14445/2020\|EPI_ISL_711772\|202 | ···················· | ···················· | ····················· | ···················· | ···················· | ···················· |
| X | 4135 | hCoV-19/Denmark/DCGC-14447/2020\|EPI_ISL_711773\|202 | ···················· | ···················· | ····················· | ···················· | ···················· | ···················· |
| X | 4136 | hCoV-19/Denmark/DCGC-14449/2020\|EPI_ISL_711774\|202 | ···················· | ···················· | ····················· | ···················· | ···················· | ···················· |
| X | 4137 | hCoV-19/Denmark/DCGC-14453/2020\|EPI_ISL_711775\|202 | ···················· | ···················· | ····················· | ···················· | ···················· | ···················· |
| X | 4138 | hCoV-19/Denmark/DCGC-14455/2020\|EPI_ISL_711776\|202 | ···················· | ···················· | ····················· | ···················· | ···················· | ···················· |
| X | 4139 | hCoV-19/Denmark/DCGC-14456/2020\|EPI_ISL_711777\|202 | ···················· | ···················· | ····················· | ···················· | ···················· | ···················· |
| X | 4140 | hCoV-19/Denmark/DCGC-14457/2020\|EPI_ISL_711778\|202 | ···················· | ···················· | ····················· | ···················· | ···················· | ···················· |
| X | 4141 | hCoV-19/Denmark/DCGC-14459/2020\|EPI_ISL_711779\|202 | ···················· | ···················· | ····················· | ···················· | ···················· | ···················· |
| X | 4142 | hCoV-19/Denmark/DCGC-14460/2020\|EPI_ISL_711780\|202 | ···················· | ···················· | ····················· | ···················· | ···················· | ···················· |
| X | 4143 | hCoV-19/Denmark/DCGC-14462/2020\|EPI_ISL_711781\|202 | ···················· | ···················· | ····················· | ···················· | ···················· | ···················· |
| X | 4144 | hCoV-19/Denmark/DCGC-14463/2020\|EPI_ISL_711782\|202 | ···················· | ···················· | ····················· | ···················· | ···················· | ···················· |
| X | 4145 | hCoV-19/Denmark/DCGC-14464/2020\|EPI_ISL_711783\|202 | ···················· | ···················· | ····················· | ···················· | ···················· | ···················· |
| X | 4146 | hCoV-19/Denmark/DCGC-14465/2020\|EPI_ISL_711784\|202 | ···················· | ···················· | ····················· | ···················· | ···················· | ···················· |
| X | 4147 | hCoV-19/Denmark/DCGC-14468/2020\|EPI_ISL_711785\|202 | ···················· | ···················· | ····················· | ···················· | ···················· | ···················· |
| X | 4148 | hCoV-19/Denmark/DCGC-14471/2020\|EPI_ISL_711786\|202 | ···················· | ···················· | ····················· | ···················· | ···················· | ···················· |
| X | 4149 | hCoV-19/Denmark/DCGC-14472/2020\|EPI_ISL_711787\|202 | ···················· | ···················· | ····················· | ···················· | ···················· | ···················· |
| X | 4150 | hCoV-19/Denmark/DCGC-14474/2020\|EPI_ISL_711788\|202 | ···················· | ···················· | ····················· | ···················· | ···················· | ···················· |
| X | 4151 | hCoV-19/Denmark/DCGC-14476/2020\|EPI_ISL_711789\|202 | ···················· | ···················· | ····················· | ···················· | ···················· | ···················· |
| X | 4152 | hCoV-19/Denmark/DCGC-14480/2020\|EPI_ISL_711790\|202 | ···················· | ···················· | ····················· | ···················· | ···················· | ···················· |
| X | 4153 | hCoV-19/Denmark/DCGC-14481/2020\|EPI_ISL_711791\|202 | ···················· | ·A·················· | ····················· | ···················· | ···················· | ···················· |
| X | 4154 | hCoV-19/Denmark/DCGC-14483/2020\|EPI_ISL_711792\|202 | ···················· | ···················· | ····················· | ···················· | ···················· | ···················· |
| X | 4155 | hCoV-19/Denmark/DCGC-14486/2020\|EPI_ISL_711793\|202 | ···················· | ···················· | ····················· | ···················· | ···················· | ···················· |
| X | 4156 | hCoV-19/Denmark/DCGC-14488/2020\|EPI_ISL_711794\|202 | ···················· | ···················· | ····················· | ···················· | ···················· | ···················· |
| X | 4157 | hCoV-19/Denmark/DCGC-14490/2020\|EPI_ISL_711795\|202 | ···················· | ·A·················· | ····················· | ···················· | ···················· | ···················· |
| X | 4158 | hCoV-19/Denmark/DCGC-14491/2020\|EPI_ISL_711796\|202 | ···················· | ···················· | ····················· | ···················· | ···················· | ···················· |
| X | 4159 | hCoV-19/Denmark/DCGC-14495/2020\|EPI_ISL_711797\|202 | ···················· | ···················· | ····················· | ···················· | ···················· | ···················· |
| X | 4160 | hCoV-19/Denmark/DCGC-14498/2020\|EPI_ISL_711798\|202 | ···················· | ···················· | ····················· | ···················· | ···················· | ···················· |
| X | 4161 | hCoV-19/Denmark/DCGC-14499/2020\|EPI_ISL_711799\|202 | ···················· | ·A·················· | ····················· | ···················· | ···················· | ···················· |
| X | 4162 | hCoV-19/England/QEUH-BA34AE/2020\|EPI_ISL_721100\|20 | ···················· | ···················· | ····················· | ···················· | ···················· | ···················· |
| X | 4163 | hCoV-19/England/QEUH-B9F8A9/2020\|EPI_ISL_721101\|20 | ···················· | ···················· | ····················· | ···················· | ···················· | ···················· |
| X | 4164 | hCoV-19/England/QEUH-B9FA49/2020\|EPI_ISL_721102\|20 | ···················· | ···················· | ····················· | ···················· | ······T············· | ···················· |
| X | 4165 | hCoV-19/England/QEUH-B9F751/2020\|EPI_ISL_721103\|20 | ···················· | ···················· | ····················· | ···················· | ···················· | ···················· |
| X | 4166 | hCoV-19/England/QEUH-BA3918/2020\|EPI_ISL_721104\|20 | ···················· | ···················· | ····················· | ···················· | ···················· | ···················· |
| X | 4167 | hCoV-19/England/QEUH-B9F311/2020\|EPI_ISL_721105\|20 | ···················· | ···················· | ····················· | ···················· | ···················· | ···················· |
| X | 4168 | hCoV-19/England/QEUH-B9FA3A/2020\|EPI_ISL_721106\|20 | ···················· | ···················· | ····················· | ···················· | ···················· | ···················· |
| X | 4169 | hCoV-19/England/QEUH-BA3277/2020\|EPI_ISL_721107\|20 | ···················· | ···················· | ····················· | ···················· | ···················· | ···················· |
| X | 4170 | hCoV-19/England/QEUH-BA349F/2020\|EPI_ISL_721108\|20 | ···················· | ···················· | ····················· | ···················· | ···················· | ···················· |
| X | 4171 | hCoV-19/England/QEUH-B9E3E5/2020\|EPI_ISL_721109\|20 | ···················· | ···················· | ····················· | ···················· | ···················· | ···················· |
| X | 4172 | hCoV-19/England/QEUH-B9F663/2020\|EPI_ISL_721110\|20 | ···················· | ···················· | ····················· | ···················· | ···················· | ···················· |
| X | 4173 | hCoV-19/England/QEUH-B9F6AF/2020\|EPI_ISL_721112\|20 | ···················· | ···················· | ····················· | ···················· | ···················· | ···················· |
| X | 4174 | hCoV-19/England/QEUH-B9F690/2020\|EPI_ISL_721113\|20 | ···················· | ···················· | ····················· | ···················· | ···················· | ···················· |
| X | 4175 | hCoV-19/England/QEUH-BA32A4/2020\|EPI_ISL_721114\|20 | ···················· | ···················· | ····················· | ···················· | ···················· | ···················· |

| | | | | | | | | |
|---|---|---|---|---|---|---|---|---|
| X | 4176 | hCoV-19/England/QEUH-BA32E0/2020\|EPI_ISL_721115\|20 | ···················· | ···················· | ···················· | ···················· | ···················· | ···················· |
| X | 4177 | hCoV-19/England/QEUH-B9E181/2020\|EPI_ISL_721119\|20 | ···················· | ···················· | ···················· | ···················· | ···················· | ···················· |
| X | 4178 | hCoV-19/England/QEUH-B9F92E/2020\|EPI_ISL_721120\|20 | ···················· | ···················· | ···················· | ···················· | ·····T·············· | ···················· |
| X | 4179 | hCoV-19/England/QEUH-B9FA94/2020\|EPI_ISL_721121\|20 | ···················· | ···················· | ···················· | ···················· | ···················· | ···················· |
| X | 4180 | hCoV-19/England/QEUH-B9F9F1/2020\|EPI_ISL_721122\|20 | ···················· | ···················· | ···················· | ···················· | ···················· | ···················· |
| X | 4181 | hCoV-19/England/QEUH-B9F575/2020\|EPI_ISL_721123\|20 | ···················· | ···················· | ···················· | ···················· | ···················· | ···················· |
| X | 4182 | hCoV-19/England/QEUH-B9FD31/2020\|EPI_ISL_721124\|20 | ···················· | ···················· | ···················· | ···················· | ···················· | ···················· |
| X | 4183 | hCoV-19/England/QEUH-BA3347/2020\|EPI_ISL_721125\|20 | ···················· | ···················· | ···················· | ···················· | ···················· | ···················· |
| X | 4184 | hCoV-19/England/QEUH-B9FD9B/2020\|EPI_ISL_721126\|20 | ···················· | ···················· | ···················· | ···················· | ···················· | ···················· |
| X | 4185 | hCoV-19/England/QEUH-BA331A/2020\|EPI_ISL_721127\|20 | ···················· | ···················· | ···················· | ···················· | ···················· | ···················· |
| X | 4186 | hCoV-19/England/QEUH-B9FBBF/2020\|EPI_ISL_721128\|20 | ···················· | ···················· | ···················· | ···················· | ···················· | ···················· |
| X | 4187 | hCoV-19/England/QEUH-B9F3B7/2020\|EPI_ISL_721132\|20 | ···················· | ···················· | ···················· | ···················· | ···················· | ···················· |
| X | 4188 | hCoV-19/England/QEUH-B9E224/2020\|EPI_ISL_721133\|20 | ···················· | ···················· | ···················· | ···················· | ···················· | ···················· |
| X | 4189 | hCoV-19/England/QEUH-B9F997/2020\|EPI_ISL_721134\|20 | ···················· | ···················· | ···················· | ···················· | ···················· | ···················· |
| X | 4190 | hCoV-19/England/QEUH-BA2D1D/2020\|EPI_ISL_721136\|20 | ···················· | ···················· | ···················· | ···················· | ···················· | ···················· |
| X | 4191 | hCoV-19/England/QEUH-B9E057/2020\|EPI_ISL_721137\|20 | ···················· | ···················· | ···················· | ···················· | ···················· | ···················· |
| X | 4192 | hCoV-19/England/QEUH-B9FAEF/2020\|EPI_ISL_721138\|20 | ···················· | ···················· | ···················· | ···················· | ···················· | ···················· |
| X | 4193 | hCoV-19/England/QEUH-B9FD22/2020\|EPI_ISL_721139\|20 | ···················· | ···················· | ···················· | ···················· | ···················· | ···················· |
| X | 4194 | hCoV-19/England/QEUH-B9F557/2020\|EPI_ISL_721140\|20 | ···················· | ···················· | ···················· | ···················· | ···················· | ···················· |
| X | 4195 | hCoV-19/England/QEUH-BA2D0E/2020\|EPI_ISL_721141\|20 | ···················· | ···················· | ···················· | ···················· | ·····T·············· | ···················· |
| X | 4196 | hCoV-19/England/QEUH-B9FAFE/2020\|EPI_ISL_721142\|20 | ···················· | ···················· | ···················· | ···················· | ···················· | ···················· |
| X | 4197 | hCoV-19/England/QEUH-BA2CE3/2020\|EPI_ISL_721144\|20 | ···················· | ···················· | ···················· | ···················· | ···················· | ···················· |
| X | 4198 | hCoV-19/England/QEUH-B9F6BE/2020\|EPI_ISL_721145\|20 | ···················· | ···················· | ···················· | ···················· | ···················· | ···················· |
| X | 4199 | hCoV-19/England/QEUH-B9E066/2020\|EPI_ISL_721146\|20 | ···················· | ···················· | ···················· | ···················· | ···················· | ···················· |
| X | 4200 | hCoV-19/England/QEUH-BA34CC/2020\|EPI_ISL_721147\|20 | ···················· | ···················· | ···················· | ···················· | ·····T·············· | ···················· |
| X | 4201 | hCoV-19/England/QEUH-B9FA2B/2020\|EPI_ISL_721148\|20 | ···················· | ···················· | ···················· | ···················· | ···················· | ···················· |
| X | 4202 | hCoV-19/England/QEUH-BA321D/2020\|EPI_ISL_721149\|20 | ···················· | ···················· | ···················· | ···················· | ···················· | ···················· |
| X | 4203 | hCoV-19/England/QEUH-B9F3F3/2020\|EPI_ISL_721150\|20 | ···················· | ···················· | ···················· | ···················· | ···················· | ···················· |
| X | 4204 | hCoV-19/England/QEUH-BA3338/2020\|EPI_ISL_721151\|20 | ···················· | ···················· | ···················· | ···················· | ···················· | ···················· |
| X | 4205 | hCoV-19/England/QEUH-BA3541/2020\|EPI_ISL_721152\|20 | ···················· | ···················· | ···················· | ···················· | ···················· | ···················· |
| X | 4206 | hCoV-19/England/QEUH-B9E0FD/2020\|EPI_ISL_721153\|20 | ···················· | ···················· | ···················· | ···················· | ···················· | ···················· |
| X | 4207 | hCoV-19/England/QEUH-B9F4B4/2020\|EPI_ISL_721154\|20 | ···················· | ···················· | ···················· | ···················· | ···················· | ···················· |
| X | 4208 | hCoV-19/England/QEUH-B9FD7D/2020\|EPI_ISL_721155\|20 | ···················· | ···················· | ···················· | ···················· | ···················· | ···················· |
| X | 4209 | hCoV-19/England/QEUH-B9F43C/2020\|EPI_ISL_721157\|20 | ···················· | ···················· | ···················· | ···················· | ···················· | ···················· |
| X | 4210 | hCoV-19/England/QEUH-BA35C9/2020\|EPI_ISL_721159\|20 | ···················· | ···················· | ···················· | ···················· | ···················· | ···················· |
| X | 4211 | hCoV-19/Netherlands/LI-EMC-122/2020\|EPI_ISL_722413 | ···················· | ···················· | ···················· | ···················· | ·····T·············· | ···················· |
| X | 4212 | hCoV-19/mink/Netherlands/NB-EMC-67-1/2020\|EPI_ISL_ | ···················· | ···················· | ···················· | ···················· | ·····T·············· | ···················· |
| X | 4213 | hCoV-19/mink/Netherlands/NB-EMC-67-18/2020\|EPI_ISL | ···················· | ···················· | ···················· | ···················· | ·····T·············· | ···················· |
| X | 4214 | hCoV-19/mink/Netherlands/NB-EMC-67-19/2020\|EPI_ISL | ···················· | ···················· | ···················· | ···················· | ·····T·············· | ···················· |
| X | 4215 | hCoV-19/mink/Netherlands/NB-EMC-67-2/2020\|EPI_ISL_ | ···················· | ···················· | ···················· | ···················· | ·····T·············· | ···················· |
| X | 4216 | hCoV-19/mink/Netherlands/NB-EMC-67-6/2020\|EPI_ISL_ | ···················· | ···················· | ···················· | ···················· | ·····T·············· | ···················· |
| X | 4217 | hCoV-19/Netherlands/ZH-EMC-981/2020\|EPI_ISL_722419 | ···················· | ···················· | ···················· | ···················· | ···················· | ···················· |
| X | 4218 | hCoV-19/Netherlands/LI-EMC-128/2020\|EPI_ISL_722420 | ···················· | ···················· | ···················· | ···················· | ·····T·············· | ···················· |
| X | 4219 | hCoV-19/Netherlands/ZH-EMC-951/2020\|EPI_ISL_722421 | ···················· | ···················· | ···················· | ···················· | ···················· | ···················· |
| X | 4220 | hCoV-19/Netherlands/ZH-EMC-964/2020\|EPI_ISL_722422 | ···················· | ···················· | ···················· | ···················· | ···················· | ···················· |
| X | 4221 | hCoV-19/Netherlands/ZH-EMC-962/2020\|EPI_ISL_722423 | ···················· | ···················· | ···················· | ···················· | ···················· | ···················· |
| X | 4222 | hCoV-19/Netherlands/UT-EMC-200/2020\|EPI_ISL_722424 | ···················· | ···················· | ···················· | ···················· | ···················· | ···················· |
| X | 4223 | hCoV-19/Netherlands/ZH-EMC-1068/2020\|EPI_ISL_72242 | ···················· | ···················· | ···················· | ···················· | ···················· | ···················· |
| X | 4224 | hCoV-19/Netherlands/un-EMC-805/2020\|EPI_ISL_722426 | ···················· | ···················· | ···················· | ···················· | ···················· | ···················· |
| X | 4225 | hCoV-19/Netherlands/un-EMC-806/2020\|EPI_ISL_722427 | ···················· | ···················· | ···················· | ···················· | ···················· | ···················· |
| X | 4226 | hCoV-19/Netherlands/un-EMC-807/2020\|EPI_ISL_722428 | ···················· | ···················· | ···················· | ···················· | ···················· | ···················· |
| X | 4227 | hCoV-19/Netherlands/un-EMC-808/2020\|EPI_ISL_722429 | ···················· | ···················· | ···················· | ···················· | ···················· | ···················· |
| X | 4228 | hCoV-19/Netherlands/un-EMC-809/2020\|EPI_ISL_722430 | ···················· | ···················· | ···················· | ···················· | ···················· | ···················· |
| X | 4229 | hCoV-19/Netherlands/un-EMC-810/2020\|EPI_ISL_722431 | ···················· | ···················· | ···················· | ···················· | ···················· | ···················· |
| X | 4230 | hCoV-19/Netherlands/un-EMC-811/2020\|EPI_ISL_722432 | ···················· | ···················· | ···················· | ···················· | ···················· | ···················· |
| X | 4231 | hCoV-19/Netherlands/ZH-EMC-1081/2020\|EPI_ISL_72243 | ···················· | ···················· | ···················· | ···················· | ···················· | ···················· |
| X | 4232 | hCoV-19/Netherlands/ZH-EMC-1056/2020\|EPI_ISL_72243 | ···················· | ···················· | ···················· | ···················· | ···················· | ···················· |
| X | 4233 | hCoV-19/Netherlands/LI-EMC-129/2020\|EPI_ISL_722437 | ···················· | ···················· | ···················· | ···················· | ·····T·············· | ···················· |
| X | 4234 | hCoV-19/Netherlands/LI-EMC-125/2020\|EPI_ISL_722438 | ···················· | ···················· | ···················· | ···················· | ·····T·············· | ···················· |
| X | 4235 | hCoV-19/Netherlands/LI-EMC-127/2020\|EPI_ISL_722439 | ···················· | ···················· | ···················· | ···················· | ·····T·············· | ···················· |
| X | 4236 | hCoV-19/Netherlands/DR-EMC-52/2020\|EPI_ISL_722440\| | ···················· | ···················· | ···················· | ···················· | ·····T·············· | ···················· |
| X | 4237 | hCoV-19/Netherlands/ZH-EMC-930/2020\|EPI_ISL_722441 | ···················· | ···················· | ···················· | ···················· | ···················· | ···················· |
| X | 4238 | hCoV-19/Netherlands/GE-EMC-380/2020\|EPI_ISL_722442 | ···················· | ·A·················· | ···················· | ···················· | ···················· | ··A················· |
| X | 4239 | hCoV-19/Netherlands/GE-EMC-419/2020\|EPI_ISL_722443 | ···················· | ···················· | ···················· | ···················· | ·····T·············· | ···················· |
| X | 4240 | hCoV-19/Netherlands/GE-EMC-384/2020\|EPI_ISL_722444 | ···················· | ·A·················· | ···················· | ···················· | ···················· | ··A················· |
| X | 4241 | hCoV-19/Netherlands/LI-EMC-123/2020\|EPI_ISL_722445 | ···················· | ···················· | ···················· | ···················· | ·····T·············· | ···················· |
| X | 4242 | hCoV-19/Netherlands/LI-EMC-124/2020\|EPI_ISL_722446 | ···················· | ···················· | ···················· | ···················· | ·····T·············· | ···················· |
| X | 4243 | hCoV-19/Netherlands/GE-EMC-388/2020\|EPI_ISL_722447 | ···················· | ·A·················· | ···················· | ···················· | ···················· | ··A················· |
| X | 4244 | hCoV-19/mink/Netherlands/NB-EMC-68-1/2020\|EPI_ISL_ | ···················· | ···················· | ···················· | ···················· | ·····T·············· | ···················· |
| X | 4245 | hCoV-19/mink/Netherlands/NB-EMC-68-14/2020\|EPI_ISL | ···················· | ···················· | ···················· | ···················· | ·····T·············· | ···················· |
| X | 4246 | hCoV-19/mink/Netherlands/NB-EMC-68-2/2020\|EPI_ISL_ | ···················· | ···················· | ···················· | ···················· | ·····T·············· | ···················· |
| X | 4247 | hCoV-19/mink/Netherlands/NB-EMC-68-3/2020\|EPI_ISL_ | ···················· | ···················· | ···················· | ···················· | ·····T·············· | ···················· |
| X | 4248 | hCoV-19/Netherlands/LI-EMC-130/2020\|EPI_ISL_722452 | ···················· | ···················· | ···················· | ···················· | ·····T·············· | ···················· |
| X | 4249 | hCoV-19/Netherlands/GE-EMC-386/2020\|EPI_ISL_722453 | ···················· | ·A·················· | ···················· | ···················· | ···················· | ··A················· |
| X | 4250 | hCoV-19/Netherlands/GE-EMC-436/2020\|EPI_ISL_722454 | ···················· | ···················· | ···················· | ···················· | ···················· | ···················· |
| X | 4251 | hCoV-19/Netherlands/NH-EMC-96/2020\|EPI_ISL_722455\| | ···················· | ···················· | ···················· | ···················· | ···················· | ···················· |
| X | 4252 | hCoV-19/Netherlands/UT-EMC-224/2020\|EPI_ISL_722456 | ···················· | ···················· | ···················· | ···················· | ···················· | ···················· |
| X | 4253 | hCoV-19/Netherlands/UT-EMC-225/2020\|EPI_ISL_722457 | ···················· | ···················· | ···················· | ···················· | ···················· | ···················· |
| X | 4254 | hCoV-19/Netherlands/UT-EMC-226/2020\|EPI_ISL_722458 | ···················· | ···················· | ···················· | ···················· | ···················· | ···················· |
| X | 4255 | hCoV-19/Netherlands/FL-EMC-45/2020\|EPI_ISL_722459\| | ···················· | ···················· | ···················· | ···················· | ···················· | ···················· |

| | | | | | | | | |
|---|---|---|---|---|---|---|---|---|
| X | 4256 | hCoV-19/Netherlands/OV-EMC-98/2020\|EPI_ISL_722463\| | ···················· | ·A·················· | ····················· | ···················· | ···················· | ···A················ |
| X | 4257 | hCoV-19/Netherlands/ZH-EMC-915/2020\|EPI_ISL_722464 | ···················· | ···················· | ····················· | ···················· | ···················· | ···················· |
| X | 4258 | hCoV-19/Netherlands/LI-EMC-121/2020\|EPI_ISL_722466 | ···········T········ | ···················· | ····················· | ···················· | ···················· | ···················· |
| X | 4259 | hCoV-19/Netherlands/NB-EMC-390/2020\|EPI_ISL_722467 | ···················· | ···················· | ····················· | ···················· | ···················· | ···················· |
| X | 4260 | hCoV-19/Netherlands/NB-EMC-391/2020\|EPI_ISL_722468 | ···················· | ···················· | ····················· | ···················· | ···················· | ···················· |
| X | 4261 | hCoV-19/Netherlands/NB-EMC-392/2020\|EPI_ISL_722469 | ···················· | ···················· | ····················· | ···················· | ···················· | ···················· |
| X | 4262 | hCoV-19/Netherlands/NB-EMC-393/2020\|EPI_ISL_722470 | ···················· | ···················· | ····················· | ···················· | ···················· | ···················· |
| X | 4263 | hCoV-19/Netherlands/NB-EMC-394/2020\|EPI_ISL_722471 | ···················· | ···················· | ····················· | ···················· | ···················· | ···················· |
| X | 4264 | hCoV-19/Netherlands/NB-EMC-395/2020\|EPI_ISL_722472 | ···················· | ···················· | ····················· | ···················· | ···················· | ···················· |
| X | 4265 | hCoV-19/Netherlands/NB-EMC-396/2020\|EPI_ISL_722473 | ···················· | ···················· | ····················· | ···················· | ···················· | ···················· |
| X | 4266 | hCoV-19/Netherlands/NB-EMC-397/2020\|EPI_ISL_722474 | ···················· | ···················· | ····················· | ···················· | ···················· | ···················· |
| X | 4267 | hCoV-19/Netherlands/NB-EMC-398/2020\|EPI_ISL_722475 | ···················· | ···················· | ····················· | ···················· | ···················· | ···················· |
| X | 4268 | hCoV-19/Netherlands/NB-EMC-399/2020\|EPI_ISL_722476 | ···················· | ·A·················· | ····················· | ···················· | ···················· | ···A················ |
| X | 4269 | hCoV-19/Netherlands/NB-EMC-400/2020\|EPI_ISL_722477 | ···················· | ···················· | ····················· | ···················· | ···················· | ···················· |
| X | 4270 | hCoV-19/Netherlands/NB-EMC-401/2020\|EPI_ISL_722478 | ···················· | ···················· | ····················· | ···················· | ···················· | ···················· |
| X | 4271 | hCoV-19/Netherlands/NB-EMC-402/2020\|EPI_ISL_722479 | ···················· | ···················· | ····················· | ···················· | ···················· | ···················· |
| X | 4272 | hCoV-19/Netherlands/NB-EMC-403/2020\|EPI_ISL_722480 | ···················· | ···················· | ····················· | ···················· | ···················· | ···················· |
| X | 4273 | hCoV-19/Netherlands/NB-EMC-404/2020\|EPI_ISL_722481 | ···················· | ···················· | ····················· | ···················· | ···················· | ···················· |
| X | 4274 | hCoV-19/Netherlands/NB-EMC-407/2020\|EPI_ISL_722482 | ···················· | ···················· | ····················· | ···················· | ···················· | ···················· |
| X | 4275 | hCoV-19/Netherlands/NB-EMC-409/2020\|EPI_ISL_722483 | ···················· | ···················· | ····················· | ···················· | ···················· | ···················· |
| X | 4276 | hCoV-19/Netherlands/NB-EMC-410/2020\|EPI_ISL_722484 | ···················· | ···················· | ····················· | ···················· | ···················· | ···················· |
| X | 4277 | hCoV-19/Netherlands/NB-EMC-412/2020\|EPI_ISL_722485 | ···················· | ···················· | ····················· | ···················· | ···················· | ···················· |
| X | 4278 | hCoV-19/Netherlands/un-EMC-802/2020\|EPI_ISL_722486 | ···················· | ···················· | ····················· | ···················· | ···················· | ···················· |
| X | 4279 | hCoV-19/Netherlands/un-EMC-803/2020\|EPI_ISL_722487 | ···················· | ···················· | ····················· | ···················· | ···················· | ···················· |
| X | 4280 | hCoV-19/Netherlands/ZH-EMC-1022/2020\|EPI_ISL_72248 | ···················· | ···················· | ····················· | ···················· | ······T············· | ···················· |
| X | 4281 | hCoV-19/Netherlands/ZH-EMC-1046/2020\|EPI_ISL_72248 | ···················· | ·A·················· | ····················· | ···················· | ···················· | ···················· |
| X | 4282 | hCoV-19/England/ALDP-B75D31/2020\|EPI_ISL_723700\|20 | ···················· | ···················· | ····················· | ···················· | ···················· | ···················· |
| X | 4283 | hCoV-19/England/ALDP-B75D6E/2020\|EPI_ISL_723701\|20 | ···················· | ···················· | ····················· | ···················· | ···················· | ···················· |
| X | 4284 | hCoV-19/Wales/ALDP-B75D8C/2020\|EPI_ISL_723702\|2020 | ···················· | ···················· | ····················· | ···················· | ···················· | ···················· |
| X | 4285 | hCoV-19/England/ALDP-B75DAA/2020\|EPI_ISL_723703\|20 | ···················· | ···················· | ····················· | ···················· | ···················· | ···················· |
| X | 4286 | hCoV-19/England/ALDP-B75DB9/2020\|EPI_ISL_723704\|20 | ···················· | ···················· | ····················· | ···················· | ···················· | ···················· |
| X | 4287 | hCoV-19/England/ALDP-B75DE6/2020\|EPI_ISL_723705\|20 | ···················· | ···················· | ····················· | ···················· | ···················· | ···················· |
| X | 4288 | hCoV-19/England/ALDP-B75DF5/2020\|EPI_ISL_723706\|20 | ···················· | ···················· | ····················· | ···················· | ···················· | ···················· |
| X | 4289 | hCoV-19/England/ALDP-B75E01/2020\|EPI_ISL_723707\|20 | ···················· | ···················· | ····················· | ···················· | ···················· | ···················· |
| X | 4290 | hCoV-19/England/ALDP-B75E10/2020\|EPI_ISL_723708\|20 | ···················· | ···················· | ····················· | ···················· | ···················· | ···················· |
| X | 4291 | hCoV-19/England/ALDP-B75E2F/2020\|EPI_ISL_723709\|20 | ···················· | ···················· | ····················· | ···················· | ···················· | ···················· |
| X | 4292 | hCoV-19/England/ALDP-B75E3E/2020\|EPI_ISL_723710\|20 | ···················· | ···················· | ····················· | ···················· | ···················· | ···················· |
| X | 4293 | hCoV-19/England/ALDP-B75E5C/2020\|EPI_ISL_723712\|20 | ···················· | ···················· | ····················· | ···················· | ···················· | ···················· |
| X | 4294 | hCoV-19/England/ALDP-B75E6B/2020\|EPI_ISL_723713\|20 | ···················· | ···················· | ····················· | ···················· | ···················· | ···················· |
| X | 4295 | hCoV-19/England/ALDP-B76028/2020\|EPI_ISL_723725\|20 | ···················· | ···················· | ····················· | ···················· | ···················· | ···················· |
| X | 4296 | hCoV-19/England/ALDP-B76064/2020\|EPI_ISL_723727\|20 | ···················· | ···················· | ····················· | ···················· | ···················· | ···················· |
| X | 4297 | hCoV-19/England/ALDP-B76073/2020\|EPI_ISL_723728\|20 | ···················· | ···················· | ····················· | ···················· | ···················· | ···················· |
| X | 4298 | hCoV-19/England/ALDP-B76125/2020\|EPI_ISL_723736\|20 | ···················· | ···················· | ····················· | ···················· | ···················· | ···················· |
| X | 4299 | hCoV-19/England/ALDP-B76134/2020\|EPI_ISL_723737\|20 | ···················· | ···················· | ····················· | ···················· | ···················· | ···················· |
| X | 4300 | hCoV-19/England/ALDP-B76143/2020\|EPI_ISL_723738\|20 | ···················· | ·A·················· | ····················· | ···················· | ···················· | ···················· |
| X | 4301 | hCoV-19/England/ALDP-B76161/2020\|EPI_ISL_723739\|20 | ···················· | ···················· | ····················· | ···················· | ···················· | ···················· |
| X | 4302 | hCoV-19/England/ALDP-B76170/2020\|EPI_ISL_723740\|20 | ···················· | ···················· | ····················· | ···················· | ···················· | ···················· |
| X | 4303 | hCoV-19/England/ALDP-B7618F/2020\|EPI_ISL_723741\|20 | ···················· | ···················· | ····················· | ···················· | ···················· | ···················· |
| X | 4304 | hCoV-19/England/ALDP-B761AD/2020\|EPI_ISL_723742\|20 | ···················· | ···················· | ····················· | ···················· | ···················· | ···················· |
| X | 4305 | hCoV-19/England/ALDP-B761BC/2020\|EPI_ISL_723743\|20 | ···················· | ···················· | ····················· | ···················· | ···················· | ···················· |
| X | 4306 | hCoV-19/England/ALDP-B761DA/2020\|EPI_ISL_723744\|20 | ···················· | ···················· | ····················· | ···················· | ···················· | ···················· |
| X | 4307 | hCoV-19/England/ALDP-B76204/2020\|EPI_ISL_723745\|20 | ···················· | ···················· | ····················· | ···················· | ···················· | ···················· |
| X | 4308 | hCoV-19/England/ALDP-B76231/2020\|EPI_ISL_723746\|20 | ···················· | ···················· | ····················· | ···················· | ···················· | ···················· |
| X | 4309 | hCoV-19/England/ALDP-B76240/2020\|EPI_ISL_723747\|20 | ···················· | ···················· | ····················· | ···················· | ···················· | ···················· |
| X | 4310 | hCoV-19/England/ALDP-B7625F/2020\|EPI_ISL_723748\|20 | ···················· | ···················· | ····················· | ···················· | ···················· | ···················· |
| X | 4311 | hCoV-19/England/ALDP-B7629B/2020\|EPI_ISL_723749\|20 | ···················· | ···················· | ····················· | ···················· | ···················· | ···················· |
| X | 4312 | hCoV-19/England/ALDP-B762AA/2020\|EPI_ISL_723750\|20 | ···················· | ···················· | ····················· | ···················· | ···················· | ···················· |
| X | 4313 | hCoV-19/England/ALDP-B762B9/2020\|EPI_ISL_723751\|20 | ···················· | ···················· | ····················· | ···················· | ···················· | ···················· |
| X | 4314 | hCoV-19/England/ALDP-B762F5/2020\|EPI_ISL_723753\|20 | ···················· | ···················· | ····················· | ···················· | ···················· | ···················· |
| X | 4315 | hCoV-19/England/ALDP-B76301/2020\|EPI_ISL_723754\|20 | ···················· | ···················· | ····················· | ···················· | ···················· | ···················· |
| X | 4316 | hCoV-19/England/ALDP-B76310/2020\|EPI_ISL_723755\|20 | ···················· | ···················· | ····················· | ···················· | ···················· | ···················· |
| X | 4317 | hCoV-19/England/ALDP-B7632F/2020\|EPI_ISL_723756\|20 | ···················· | ···················· | ····················· | ···················· | ···················· | ···················· |
| X | 4318 | hCoV-19/England/ALDP-B7633E/2020\|EPI_ISL_723757\|20 | ···················· | ···················· | ····················· | ···················· | ···················· | ···················· |
| X | 4319 | hCoV-19/England/ALDP-B7635C/2020\|EPI_ISL_723758\|20 | ···················· | ···················· | ····················· | ···················· | ···················· | ···················· |
| X | 4320 | hCoV-19/England/ALDP-B7636B/2020\|EPI_ISL_723759\|20 | ···················· | ···················· | ····················· | ···················· | ···················· | ···················· |
| X | 4321 | hCoV-19/England/ALDP-B76398/2020\|EPI_ISL_723760\|20 | ···················· | ···················· | ····················· | ···················· | ···················· | ···················· |
| X | 4322 | hCoV-19/England/ALDP-B7642C/2020\|EPI_ISL_723761\|20 | ···················· | ···················· | ····················· | ···················· | ···················· | ···················· |
| X | 4323 | hCoV-19/England/ALDP-B7643B/2020\|EPI_ISL_723762\|20 | ···················· | ···················· | ····················· | ···················· | ···················· | ···················· |
| X | 4324 | hCoV-19/England/ALDP-B76477/2020\|EPI_ISL_723763\|20 | ···················· | ···················· | ····················· | ···················· | ···················· | ···················· |
| X | 4325 | hCoV-19/England/ALDP-B76486/2020\|EPI_ISL_723764\|20 | ···················· | ···················· | ····················· | ···················· | ···················· | ···················· |
| X | 4326 | hCoV-19/England/ALDP-B764E0/2020\|EPI_ISL_723765\|20 | ···················· | ···················· | ····················· | ···················· | ···················· | ···················· |
| X | 4327 | hCoV-19/England/ALDP-B764FF/2020\|EPI_ISL_723766\|20 | ···················· | ···················· | ····················· | ···················· | ······T············· | ···················· |
| X | 4328 | hCoV-19/England/ALDP-B76538/2020\|EPI_ISL_723768\|20 | ···················· | ···················· | ····················· | ···················· | ···················· | ···················· |
| X | 4329 | hCoV-19/England/ALDP-B76556/2020\|EPI_ISL_723769\|20 | ···················· | ···················· | ····················· | ···················· | ···················· | ···················· |
| X | 4330 | hCoV-19/England/ALDP-B76592/2020\|EPI_ISL_723770\|20 | ···················· | ···················· | ····················· | ···················· | ···················· | ···················· |
| X | 4331 | hCoV-19/England/ALDP-B766DB/2020\|EPI_ISL_723772\|20 | ···················· | ···················· | ····················· | ···················· | ···················· | ···················· |
| X | 4332 | hCoV-19/England/ALDP-B766EA/2020\|EPI_ISL_723773\|20 | ···················· | ···················· | ····················· | ···················· | ···················· | ···················· |
| X | 4333 | hCoV-19/England/ALDP-B766F9/2020\|EPI_ISL_723774\|20 | ···················· | ···················· | ····················· | ···················· | ···················· | ···················· |
| X | 4334 | hCoV-19/England/ALDP-B76705/2020\|EPI_ISL_723775\|20 | ···················· | ···················· | ····················· | ···················· | ···················· | ···················· |
| X | 4335 | hCoV-19/England/ALDP-B76723/2020\|EPI_ISL_723776\|20 | ···················· | ···················· | ····················· | ···················· | ···················· | ···················· |

| | | | | | | | | |
|---|---|---|---|---|---|---|---|---|
| X | 4336 | hCoV-19/England/ALDP-B76741/2020\|EPI_ISL_723777\|20 | ···················· | ···················· | ····················· | ···················· | ···················· | ···················· |
| X | 4337 | hCoV-19/England/ALDP-B7676F/2020\|EPI_ISL_723778\|20 | ···················· | ···················· | ····················· | ···················· | ···················· | ···················· |
| X | 4338 | hCoV-19/England/ALDP-B7677E/2020\|EPI_ISL_723779\|20 | ···················· | ···················· | ····················· | ···················· | ···················· | ···················· |
| X | 4339 | hCoV-19/England/ALDP-B768B7/2020\|EPI_ISL_723780\|20 | ···················· | ···················· | ····················· | ···················· | ···················· | ···················· |
| X | 4340 | hCoV-19/England/ALDP-B768D5/2020\|EPI_ISL_723781\|20 | ···················· | ···················· | ····················· | ···················· | ···················· | ···················· |
| X | 4341 | hCoV-19/England/ALDP-B768E4/2020\|EPI_ISL_723782\|20 | ···················· | ···················· | ····················· | ···················· | ···················· | ···················· |
| X | 4342 | hCoV-19/England/ALDP-B768F3/2020\|EPI_ISL_723783\|20 | ···················· | ···················· | ····················· | ···················· | ···················· | ···················· |
| X | 4343 | hCoV-19/England/ALDP-B7690F/2020\|EPI_ISL_723784\|20 | ···················· | ···················· | ····················· | ···················· | ···················· | ···················· |
| X | 4344 | hCoV-19/England/ALDP-B7691E/2020\|EPI_ISL_723785\|20 | ···················· | ···················· | ····················· | ···················· | ···················· | ···················· |
| X | 4345 | hCoV-19/England/ALDP-B7694B/2020\|EPI_ISL_723786\|20 | ···················· | ···················· | ····················· | ···················· | ···················· | ···················· |
| X | 4346 | hCoV-19/England/ALDP-B7695A/2020\|EPI_ISL_723787\|20 | ···················· | ···················· | ····················· | ···················· | ···················· | ···················· |
| X | 4347 | hCoV-19/England/ALDP-B76969/2020\|EPI_ISL_723788\|20 | ···················· | ···················· | ····················· | ···················· | ···················· | ···················· |
| X | 4348 | hCoV-19/England/ALDP-B76987/2020\|EPI_ISL_723789\|20 | ···················· | ···················· | ····················· | ···················· | ···················· | ···················· |
| X | 4349 | hCoV-19/England/ALDP-B769E1/2020\|EPI_ISL_723795\|20 | ···················· | ···················· | ····················· | ···················· | ···················· | ···················· |
| X | 4350 | hCoV-19/England/ALDP-B76A0C/2020\|EPI_ISL_723797\|20 | ···················· | ···················· | ····················· | ···················· | ···················· | ···················· |
| X | 4351 | hCoV-19/England/ALDP-B76A39/2020\|EPI_ISL_723798\|20 | ···················· | ···················· | ····················· | ···················· | ···················· | ···················· |
| X | 4352 | hCoV-19/England/ALDP-B76A57/2020\|EPI_ISL_723799\|20 | ···················· | ···················· | ····················· | ···················· | ···················· | ···················· |
| X | 4353 | hCoV-19/Denmark/DCGC-14501/2020\|EPI_ISL_711800\|202 | ···················· | ···················· | ····················· | ···················· | ···················· | ···················· |
| X | 4354 | hCoV-19/Denmark/DCGC-14516/2020\|EPI_ISL_711812\|202 | ···················· | ···················· | ····················· | ···················· | ···················· | ···················· |
| X | 4355 | hCoV-19/Denmark/DCGC-14517/2020\|EPI_ISL_711813\|202 | ···················· | ···················· | ····················· | ···················· | ···················· | ···················· |
| X | 4356 | hCoV-19/Denmark/DCGC-14518/2020\|EPI_ISL_711814\|202 | ···················· | ···················· | ····················· | ···················· | ···················· | ···················· |
| X | 4357 | hCoV-19/Denmark/DCGC-14519/2020\|EPI_ISL_711815\|202 | ···················· | ···················· | ····················· | ···················· | ···················· | ···················· |
| X | 4358 | hCoV-19/Denmark/DCGC-14522/2020\|EPI_ISL_711816\|202 | ···················· | ····A··············· | ····················· | ···················· | ···················· | ···················· |
| X | 4359 | hCoV-19/Denmark/DCGC-14523/2020\|EPI_ISL_711817\|202 | ···················· | ···················· | ····················· | ···················· | ···················· | ···················· |
| X | 4360 | hCoV-19/Denmark/DCGC-14525/2020\|EPI_ISL_711818\|202 | ···················· | ···················· | ····················· | ···················· | ···················· | ···················· |
| X | 4361 | hCoV-19/Denmark/DCGC-14527/2020\|EPI_ISL_711819\|202 | ···················· | ···················· | ····················· | ···················· | ···················· | ···················· |
| X | 4362 | hCoV-19/Denmark/DCGC-14530/2020\|EPI_ISL_711820\|202 | ···················· | ···················· | ····················· | ···················· | ···················· | ···················· |
| X | 4363 | hCoV-19/Denmark/DCGC-14532/2020\|EPI_ISL_711821\|202 | ···················· | ···················· | ····················· | ···················· | ···················· | ···················· |
| X | 4364 | hCoV-19/Denmark/DCGC-14534/2020\|EPI_ISL_711822\|202 | ···················· | ·A·················· | ····················· | ···················· | ···················· | ···A················ |
| X | 4365 | hCoV-19/Denmark/DCGC-14535/2020\|EPI_ISL_711823\|202 | ···················· | ·A·················· | ····················· | ···················· | ···················· | ···A················ |
| X | 4366 | hCoV-19/Denmark/DCGC-14537/2020\|EPI_ISL_711824\|202 | ···················· | ···················· | ····················· | ···················· | ···················· | ···················· |
| X | 4367 | hCoV-19/Denmark/DCGC-14539/2020\|EPI_ISL_711825\|202 | ···················· | ···················· | ····················· | ···················· | ···················· | ···················· |
| X | 4368 | hCoV-19/Denmark/DCGC-14540/2020\|EPI_ISL_711826\|202 | ···················· | ···················· | ····················· | ···················· | ···················· | ···················· |
| X | 4369 | hCoV-19/Denmark/DCGC-14541/2020\|EPI_ISL_711827\|202 | ···················· | ···················· | ····················· | ···················· | ···················· | ···················· |
| X | 4370 | hCoV-19/Denmark/DCGC-14542/2020\|EPI_ISL_711828\|202 | ···················· | ·A·················· | ····················· | ···················· | ···················· | ···A················ |
| X | 4371 | hCoV-19/Denmark/DCGC-14544/2020\|EPI_ISL_711829\|202 | ···················· | ···················· | ····················· | ···················· | ···················· | ···················· |
| X | 4372 | hCoV-19/Denmark/DCGC-14547/2020\|EPI_ISL_711830\|202 | ···················· | ···················· | ····················· | ···················· | ···················· | ···················· |
| X | 4373 | hCoV-19/Denmark/DCGC-14548/2020\|EPI_ISL_711831\|202 | ···················· | ···················· | ····················· | ···················· | ···················· | ···················· |
| X | 4374 | hCoV-19/Denmark/DCGC-14550/2020\|EPI_ISL_711832\|202 | ···················· | ···················· | ····················· | ···················· | ···················· | ···················· |
| X | 4375 | hCoV-19/Denmark/DCGC-14551/2020\|EPI_ISL_711833\|202 | ···················· | ···················· | ····················· | ···················· | ···················· | ···················· |
| X | 4376 | hCoV-19/Denmark/DCGC-15753/2020\|EPI_ISL_714000\|202 | ···················· | ···················· | ····················· | ···················· | ···················· | ···················· |
| X | 4377 | hCoV-19/Denmark/DCGC-15754/2020\|EPI_ISL_714001\|202 | ···················· | ···················· | ····················· | ···················· | ···················· | ···················· |
| X | 4378 | hCoV-19/Denmark/DCGC-15755/2020\|EPI_ISL_714002\|202 | ···················· | ···················· | ····················· | ···················· | ···················· | ···················· |
| X | 4379 | hCoV-19/Denmark/DCGC-15756/2020\|EPI_ISL_714003\|202 | ···················· | ···················· | ····················· | ···················· | ···················· | ···················· |
| X | 4380 | hCoV-19/Denmark/DCGC-15758/2020\|EPI_ISL_714004\|202 | ···················· | ···················· | ····················· | ···················· | ···················· | ···················· |
| X | 4381 | hCoV-19/Denmark/DCGC-15761/2020\|EPI_ISL_714005\|202 | ···················· | ···················· | ····················· | ···················· | ···················· | ···················· |
| X | 4382 | hCoV-19/Denmark/DCGC-15762/2020\|EPI_ISL_714006\|202 | ···················· | ···················· | ····················· | ···················· | ···················· | ···················· |
| X | 4383 | hCoV-19/Denmark/DCGC-15763/2020\|EPI_ISL_714007\|202 | ···················· | ···················· | ····················· | ···················· | ···················· | ···················· |
| X | 4384 | hCoV-19/Denmark/DCGC-15766/2020\|EPI_ISL_714008\|202 | ···················· | ···················· | ····················· | ···················· | ···················· | ···················· |
| X | 4385 | hCoV-19/Denmark/DCGC-15767/2020\|EPI_ISL_714009\|202 | ···················· | ···················· | ····················· | ···················· | ···················· | ···················· |
| X | 4386 | hCoV-19/Denmark/DCGC-15769/2020\|EPI_ISL_714010\|202 | ···················· | ···················· | ····················· | ···················· | ···················· | ···················· |
| X | 4387 | hCoV-19/Denmark/DCGC-15770/2020\|EPI_ISL_714011\|202 | ···················· | ·A·················· | ····················· | ···················· | ···················· | ···A················ |
| X | 4388 | hCoV-19/Denmark/DCGC-15771/2020\|EPI_ISL_714012\|202 | ···················· | ···················· | ····················· | ···················· | ···················· | ···················· |
| X | 4389 | hCoV-19/Denmark/DCGC-15776/2020\|EPI_ISL_714013\|202 | ···················· | ···················· | ····················· | ···················· | ···················· | ···················· |
| X | 4390 | hCoV-19/Denmark/DCGC-15778/2020\|EPI_ISL_714014\|202 | ···················· | ···················· | ····················· | ···················· | ···················· | ···················· |
| X | 4391 | hCoV-19/Denmark/DCGC-15779/2020\|EPI_ISL_714015\|202 | ···················· | ···················· | ····················· | ···················· | ···················· | ···················· |
| X | 4392 | hCoV-19/Denmark/DCGC-15782/2020\|EPI_ISL_714016\|202 | ···················· | ···················· | ····················· | ···················· | ···················· | ···················· |
| X | 4393 | hCoV-19/Denmark/DCGC-15785/2020\|EPI_ISL_714017\|202 | ···················· | ···················· | ····················· | ···················· | ······T············· | ···················· |
| X | 4394 | hCoV-19/Denmark/DCGC-15787/2020\|EPI_ISL_714018\|202 | ···················· | ····A··············· | ····················· | ···················· | ···················· | ···················· |
| X | 4395 | hCoV-19/Denmark/DCGC-15788/2020\|EPI_ISL_714019\|202 | ···················· | ···················· | ····················· | ···················· | ···················· | ···················· |
| X | 4396 | hCoV-19/Denmark/DCGC-15789/2020\|EPI_ISL_714020\|202 | ···················· | ···················· | ····················· | ···················· | ···················· | ···················· |
| X | 4397 | hCoV-19/Denmark/DCGC-15790/2020\|EPI_ISL_714021\|202 | ···················· | ···················· | ····················· | ···················· | ···················· | ···················· |
| X | 4398 | hCoV-19/Denmark/DCGC-15791/2020\|EPI_ISL_714022\|202 | ···················· | ···················· | ····················· | ···················· | ······T············· | ···················· |
| X | 4399 | hCoV-19/Denmark/DCGC-15793/2020\|EPI_ISL_714023\|202 | ···················· | ···················· | ····················· | ···················· | ···················· | ···················· |
| X | 4400 | hCoV-19/Denmark/DCGC-15794/2020\|EPI_ISL_714024\|202 | ···················· | ···················· | ····················· | ···················· | ···················· | ···················· |
| X | 4401 | hCoV-19/Denmark/DCGC-15795/2020\|EPI_ISL_714025\|202 | ···················· | ···················· | ····················· | ···················· | ···················· | ···················· |
| X | 4402 | hCoV-19/Denmark/DCGC-15796/2020\|EPI_ISL_714026\|202 | ···················· | ···················· | ····················· | ···················· | ···················· | ···················· |
| X | 4403 | hCoV-19/Denmark/DCGC-15798/2020\|EPI_ISL_714027\|202 | ···················· | ·A·················· | ····················· | ···················· | ···················· | ···A················ |
| X | 4404 | hCoV-19/Denmark/DCGC-15801/2020\|EPI_ISL_714028\|202 | ···················· | ···················· | ····················· | ···················· | ···················· | ···················· |
| X | 4405 | hCoV-19/Denmark/DCGC-15803/2020\|EPI_ISL_714029\|202 | ···················· | ···················· | ····················· | ···················· | ···················· | ···················· |
| X | 4406 | hCoV-19/Denmark/DCGC-15804/2020\|EPI_ISL_714030\|202 | ···················· | ·A·················· | ····················· | ···················· | ···················· | ···················· |
| X | 4407 | hCoV-19/Denmark/DCGC-15805/2020\|EPI_ISL_714031\|202 | ···················· | ···················· | ····················· | ···················· | ···················· | ···················· |
| X | 4408 | hCoV-19/Denmark/DCGC-15807/2020\|EPI_ISL_714032\|202 | ···················· | ···················· | ····················· | ···················· | ···················· | ···················· |
| X | 4409 | hCoV-19/Denmark/DCGC-15808/2020\|EPI_ISL_714033\|202 | ···················· | ··········A········· | ····················· | ···················· | ···················· | ···················· |
| X | 4410 | hCoV-19/Denmark/DCGC-15809/2020\|EPI_ISL_714034\|202 | ···················· | ···················· | ····················· | ···················· | ···················· | ···················· |
| X | 4411 | hCoV-19/Denmark/DCGC-15810/2020\|EPI_ISL_714035\|202 | ···················· | ···················· | ····················· | ···················· | ···················· | ···················· |
| X | 4412 | hCoV-19/Denmark/DCGC-15812/2020\|EPI_ISL_714036\|202 | ···················· | ···················· | ····················· | ···················· | ···················· | ···················· |
| X | 4413 | hCoV-19/Denmark/DCGC-15813/2020\|EPI_ISL_714037\|202 | ···················· | ·A·················· | ····················· | ···················· | ···················· | ···A················ |
| X | 4414 | hCoV-19/Denmark/DCGC-15815/2020\|EPI_ISL_714038\|202 | ···················· | ···················· | ····················· | ···················· | ···················· | ···················· |
| X | 4415 | hCoV-19/Denmark/DCGC-15817/2020\|EPI_ISL_714039\|202 | ···················· | ···················· | ····················· | ···················· | ···················· | ···················· |

| | | | | | | | | |
|---|---|---|---|---|---|---|---|---|
| X | 4416 | hCoV-19/Denmark/DCGC-15818/2020\|EPI_ISL_714040\|202 | ···················· | ···················· | ····················· | ···················· | ···················· | ···················· |
| X | 4417 | hCoV-19/Denmark/DCGC-15820/2020\|EPI_ISL_714041\|202 | ···················· | ···················· | ····················· | ···················· | ···················· | ···················· |
| X | 4418 | hCoV-19/Denmark/DCGC-15823/2020\|EPI_ISL_714042\|202 | ···················· | ···················· | ····················· | ···················· | ···················· | ···················· |
| X | 4419 | hCoV-19/Denmark/DCGC-15826/2020\|EPI_ISL_714043\|202 | ···················· | ···················· | ····················· | ···················· | ···················· | ···················· |
| X | 4420 | hCoV-19/Denmark/DCGC-15827/2020\|EPI_ISL_714044\|202 | ···················· | ···················· | ····················· | ···················· | ···················· | ···················· |
| X | 4421 | hCoV-19/Denmark/DCGC-15828/2020\|EPI_ISL_714045\|202 | ···················· | ···················· | ····················· | ···················· | ···················· | ···················· |
| X | 4422 | hCoV-19/Denmark/DCGC-15829/2020\|EPI_ISL_714046\|202 | ···················· | ···················· | ····················· | ···················· | ···················· | ···················· |
| X | 4423 | hCoV-19/Denmark/DCGC-15830/2020\|EPI_ISL_714047\|202 | ···················· | ···················· | ····················· | ···················· | ···················· | ···················· |
| X | 4424 | hCoV-19/Denmark/DCGC-15832/2020\|EPI_ISL_714048\|202 | ···················· | ···················· | ····················· | ···················· | ···················· | ···················· |
| X | 4425 | hCoV-19/Denmark/DCGC-15833/2020\|EPI_ISL_714049\|202 | ···················· | ···················· | ····················· | ···················· | ···················· | ···················· |
| X | 4426 | hCoV-19/Denmark/DCGC-15834/2020\|EPI_ISL_714050\|202 | ···················· | ···················· | ····················· | ···················· | ···················· | ···················· |
| X | 4427 | hCoV-19/Denmark/DCGC-15835/2020\|EPI_ISL_714051\|202 | ···················· | ···················· | ····················· | ···················· | ···················· | ···················· |
| X | 4428 | hCoV-19/Denmark/DCGC-15837/2020\|EPI_ISL_714052\|202 | ···················· | ···················· | ····················· | ···················· | ···················· | ···················· |
| X | 4429 | hCoV-19/Denmark/DCGC-15839/2020\|EPI_ISL_714053\|202 | ···················· | ···················· | ····················· | ···················· | ···················· | ···················· |
| X | 4430 | hCoV-19/Denmark/DCGC-15840/2020\|EPI_ISL_714054\|202 | ···················· | ···················· | ····················· | ···················· | ···················· | ···················· |
| X | 4431 | hCoV-19/Denmark/DCGC-15842/2020\|EPI_ISL_714055\|202 | ···················· | ···················· | ····················· | ···················· | ···················· | ···················· |
| X | 4432 | hCoV-19/Denmark/DCGC-15844/2020\|EPI_ISL_714056\|202 | ···················· | ·A·················· | ····················· | ···················· | ···················· | ···A················ |
| X | 4433 | hCoV-19/Denmark/DCGC-15846/2020\|EPI_ISL_714057\|202 | ···················· | ···················· | ····················· | ···················· | ···················· | ···················· |
| X | 4434 | hCoV-19/Denmark/DCGC-15847/2020\|EPI_ISL_714058\|202 | ···················· | ···················· | ····················· | ···················· | ···················· | ···················· |
| X | 4435 | hCoV-19/Denmark/DCGC-15850/2020\|EPI_ISL_714059\|202 | ···················· | ···················· | ····················· | ···················· | ···················· | ···················· |
| X | 4436 | hCoV-19/Denmark/DCGC-15851/2020\|EPI_ISL_714060\|202 | ···················· | ···················· | ····················· | ···················· | ···················· | ···················· |
| X | 4437 | hCoV-19/Denmark/DCGC-15852/2020\|EPI_ISL_714061\|202 | ···················· | ···················· | ····················· | ···················· | ···················· | ···················· |
| X | 4438 | hCoV-19/Denmark/DCGC-15853/2020\|EPI_ISL_714062\|202 | ···················· | ···················· | ····················· | ···················· | ···················· | ···················· |
| X | 4439 | hCoV-19/Denmark/DCGC-15854/2020\|EPI_ISL_714063\|202 | ···················· | ···················· | ····················· | ···················· | ···················· | ···················· |
| X | 4440 | hCoV-19/Denmark/DCGC-15855/2020\|EPI_ISL_714064\|202 | ···················· | ·A·················· | ····················· | ···················· | ···················· | ···A················ |
| X | 4441 | hCoV-19/Denmark/DCGC-15856/2020\|EPI_ISL_714065\|202 | ···················· | ···················· | ····················· | ···················· | ···················· | ···················· |
| X | 4442 | hCoV-19/Denmark/DCGC-15857/2020\|EPI_ISL_714066\|202 | ···················· | ···················· | ····················· | ···················· | ······T············· | ···················· |
| X | 4443 | hCoV-19/Denmark/DCGC-15859/2020\|EPI_ISL_714067\|202 | ···················· | ···················· | ····················· | ···················· | ···················· | ···················· |
| X | 4444 | hCoV-19/Denmark/DCGC-15860/2020\|EPI_ISL_714068\|202 | ···················· | ···················· | ····················· | ···················· | ···················· | ···················· |
| X | 4445 | hCoV-19/Denmark/DCGC-15862/2020\|EPI_ISL_714069\|202 | ···················· | ···················· | ····················· | ···················· | ···················· | ···················· |
| X | 4446 | hCoV-19/Denmark/DCGC-15863/2020\|EPI_ISL_714070\|202 | ···················· | ···················· | ····················· | ···················· | ···················· | ···················· |
| X | 4447 | hCoV-19/Denmark/DCGC-15864/2020\|EPI_ISL_714071\|202 | ···················· | ···················· | ····················· | ···················· | ···················· | ···················· |
| X | 4448 | hCoV-19/Denmark/DCGC-15866/2020\|EPI_ISL_714072\|202 | ···················· | ·A·················· | ····················· | ···················· | ···················· | ···················· |
| X | 4449 | hCoV-19/Denmark/DCGC-15874/2020\|EPI_ISL_714078\|202 | ···················· | ···················· | ····················· | ···················· | ···················· | ···················· |
| X | 4450 | hCoV-19/Denmark/DCGC-15875/2020\|EPI_ISL_714079\|202 | ···················· | ···················· | ····················· | ···················· | ···················· | ···················· |
| X | 4451 | hCoV-19/Denmark/DCGC-15895/2020\|EPI_ISL_714090\|202 | ···················· | ···················· | ····················· | ···················· | ···················· | ···················· |
| X | 4452 | hCoV-19/Denmark/DCGC-15897/2020\|EPI_ISL_714091\|202 | ···················· | ·A·················· | ····················· | ···················· | ···················· | ···A················ |
| X | 4453 | hCoV-19/Denmark/DCGC-15901/2020\|EPI_ISL_714092\|202 | ···················· | ···················· | ····················· | ···················· | ···················· | ···················· |
| X | 4454 | hCoV-19/Denmark/DCGC-15902/2020\|EPI_ISL_714093\|202 | ···················· | ·A·················· | ····················· | ···················· | ···················· | ···················· |
| X | 4455 | hCoV-19/Denmark/DCGC-15903/2020\|EPI_ISL_714094\|202 | ···················· | ···················· | ····················· | ···················· | ···················· | ···················· |
| X | 4456 | hCoV-19/Denmark/DCGC-15905/2020\|EPI_ISL_714095\|202 | ···················· | ···················· | ····················· | ···················· | ···················· | ···················· |
| X | 4457 | hCoV-19/Denmark/DCGC-15907/2020\|EPI_ISL_714096\|202 | ···················· | ···················· | ····················· | ···················· | ···················· | ···················· |
| X | 4458 | hCoV-19/Denmark/DCGC-15908/2020\|EPI_ISL_714097\|202 | ···················· | ···················· | ····················· | ···················· | ···················· | ···················· |
| X | 4459 | hCoV-19/Brazil/RJ-00507/2020\|EPI_ISL_717900\|2020-0 | ···················· | ···················· | ····················· | ···················· | ···················· | ···················· |
| X | 4460 | hCoV-19/Brazil/RJ-00508/2020\|EPI_ISL_717901\|2020-0 | ···················· | ···················· | ····················· | ···················· | ···················· | ···················· |
| X | 4461 | hCoV-19/Brazil/RJ-00511/2020\|EPI_ISL_717902\|2020-0 | ···················· | ···················· | ····················· | ···················· | ···················· | ···················· |
| X | 4462 | hCoV-19/Brazil/RJ-00512/2020\|EPI_ISL_717903\|2020-0 | ···················· | ···················· | ····················· | ···················· | ···················· | ···················· |
| X | 4463 | hCoV-19/Brazil/RJ-00515/2020\|EPI_ISL_717904\|2020-0 | ···················· | ···················· | ····················· | ···················· | ···················· | ···················· |
| X | 4464 | hCoV-19/Brazil/RJ-00516/2020\|EPI_ISL_717905\|2020-0 | ···················· | ···················· | ····················· | ···················· | ···················· | ···················· |
| X | 4465 | hCoV-19/Brazil/RJ-00517/2020\|EPI_ISL_717906\|2020-0 | ···················· | ···················· | ····················· | ···················· | ···················· | ···················· |
| X | 4466 | hCoV-19/Brazil/RJ-00518/2020\|EPI_ISL_717907\|2020-0 | ···················· | ···················· | ····················· | ···················· | ···················· | ···················· |
| X | 4467 | hCoV-19/Brazil/RJ-00520/2020\|EPI_ISL_717908\|2020-0 | ···················· | ···················· | ····················· | ···················· | ···················· | ···················· |
| X | 4468 | hCoV-19/Brazil/RJ-00522/2020\|EPI_ISL_717909\|2020-0 | ···················· | ···················· | ····················· | ···················· | ···················· | ···················· |
| X | 4469 | hCoV-19/Brazil/RJ-00524/2020\|EPI_ISL_717910\|2020-0 | ···················· | ···················· | ····················· | ···················· | ···················· | ···················· |
| X | 4470 | hCoV-19/Brazil/RJ-00525/2020\|EPI_ISL_717911\|2020-0 | ···················· | ···················· | ····················· | ···················· | ···················· | ···················· |
| X | 4471 | hCoV-19/Brazil/RJ-00526/2020\|EPI_ISL_717912\|2020-0 | ···················· | ···················· | ····················· | ···················· | ···················· | ···················· |
| X | 4472 | hCoV-19/Brazil/RJ-00527/2020\|EPI_ISL_717913\|2020-0 | ···················· | ···················· | ····················· | ···················· | ···················· | ···················· |
| X | 4473 | hCoV-19/Brazil/RJ-00528/2020\|EPI_ISL_717914\|2020-0 | ···················· | ···················· | ····················· | ···················· | ···················· | ···················· |
| X | 4474 | hCoV-19/Brazil/RJ-00529/2020\|EPI_ISL_717915\|2020-0 | ···················· | ···················· | ····················· | ···················· | ···················· | ···················· |
| X | 4475 | hCoV-19/Brazil/RJ-00530/2020\|EPI_ISL_717916\|2020-0 | ···················· | ···················· | ····················· | ···················· | ···················· | ···················· |
| X | 4476 | hCoV-19/Brazil/RJ-00531/2020\|EPI_ISL_717917\|2020-0 | ···················· | ···················· | ····················· | ···················· | ···················· | ···················· |
| X | 4477 | hCoV-19/Brazil/RJ-00532/2020\|EPI_ISL_717918\|2020-0 | ···················· | ···················· | ····················· | ···················· | ···················· | ···················· |
| X | 4478 | hCoV-19/Brazil/RJ-00533/2020\|EPI_ISL_717919\|2020-0 | ···················· | ···················· | ····················· | ···················· | ···················· | ···················· |
| X | 4479 | hCoV-19/Brazil/RJ-00535/2020\|EPI_ISL_717920\|2020-1 | ···················· | ···················· | ····················· | ···················· | ···················· | ···················· |
| X | 4480 | hCoV-19/Brazil/RJ-00536/2020\|EPI_ISL_717921\|2020-1 | ···················· | ···················· | ····················· | ···················· | ···················· | ···················· |
| X | 4481 | hCoV-19/Brazil/RJ-00537/2020\|EPI_ISL_717922\|2020-1 | ···················· | ···················· | ····················· | ···················· | ···················· | ···················· |
| X | 4483 | hCoV-19/Brazil/RJ-00540/2020\|EPI_ISL_717924\|2020-1 | ···················· | ···················· | ····················· | ···················· | ···················· | ···················· |
| X | 4485 | hCoV-19/Brazil/RJ-00542/2020\|EPI_ISL_717926\|2020-1 | ···················· | ···················· | ····················· | ···················· | ···················· | ···················· |
| X | 4486 | hCoV-19/Brazil/RJ-00543/2020\|EPI_ISL_717927\|2020-1 | ···················· | ···················· | ····················· | ···················· | ···················· | ···················· |
| X | 4487 | hCoV-19/Brazil/RJ-00544/2020\|EPI_ISL_717928\|2020-1 | ···················· | ···················· | ····················· | ···················· | ···················· | ···················· |
| X | 4488 | hCoV-19/Brazil/RJ-00545/2020\|EPI_ISL_717929\|2020-1 | ···················· | ···················· | ····················· | ···················· | ···················· | ···················· |
| X | 4489 | hCoV-19/Brazil/RJ-00546/2020\|EPI_ISL_717930\|2020-1 | ···················· | ···················· | ····················· | ···················· | ···················· | ···················· |
| X | 4490 | hCoV-19/Brazil/RJ-00547/2020\|EPI_ISL_717931\|2020-1 | ···················· | ···················· | ····················· | ···················· | ···················· | ···················· |
| X | 4491 | hCoV-19/Brazil/RJ-00548/2020\|EPI_ISL_717932\|2020-1 | ···················· | ···················· | ····················· | ···················· | ···················· | ···················· |
| X | 4492 | hCoV-19/Brazil/RJ-00549/2020\|EPI_ISL_717933\|2020-1 | ···················· | ···················· | ····················· | ···················· | ···················· | ···················· |
| X | 4493 | hCoV-19/Brazil/RJ-00550/2020\|EPI_ISL_717934\|2020-1 | ···················· | ···················· | ····················· | ···················· | ···················· | ···················· |
| X | 4494 | hCoV-19/Brazil/RJ-00551/2020\|EPI_ISL_717935\|2020-1 | ···················· | ···················· | ····················· | ···················· | ···················· | ···················· |
| X | 4495 | hCoV-19/Brazil/RJ-00552/2020\|EPI_ISL_717936\|2020-1 | ···················· | ···················· | ····················· | ···················· | ···················· | ···················· |
| X | 4496 | hCoV-19/Brazil/RJ-00553/2020\|EPI_ISL_717937\|2020-1 | ···················· | ···················· | ····················· | ···················· | ···················· | ···················· |
| X | 4497 | hCoV-19/Brazil/RJ-00554/2020\|EPI_ISL_717938\|2020-1 | ···················· | ···················· | ····················· | ···················· | ···················· | ···················· |

| | | | | | | | | |
|---|---|---|---|---|---|---|---|---|
| X | 4498 | hCoV-19/Brazil/RJ-00555/2020\|EPI_ISL_717939\|2020-1 | ···················· | ···················· | ····················· | ···················· | ···················· | ···················· |
| X | 4499 | hCoV-19/Brazil/RJ-00556/2020\|EPI_ISL_717940\|2020-1 | ···················· | ···················· | ····················· | ···················· | ···················· | ···················· |
| X | 4500 | hCoV-19/Brazil/RJ-00557/2020\|EPI_ISL_717941\|2020-1 | ···················· | ···················· | ····················· | ···················· | ···················· | ···················· |
| X | 4501 | hCoV-19/Brazil/RJ-00558/2020\|EPI_ISL_717942\|2020-1 | ···················· | ···················· | ····················· | ···················· | ···················· | ···················· |
| X | 4502 | hCoV-19/Brazil/RJ-00559/2020\|EPI_ISL_717943\|2020-1 | ···················· | ···················· | ····················· | ···················· | ···················· | ···················· |
| X | 4503 | hCoV-19/Brazil/RJ-00560/2020\|EPI_ISL_717944\|2020-1 | ···················· | ···················· | ····················· | ···················· | ···················· | ···················· |
| X | 4504 | hCoV-19/Brazil/RJ-00561/2020\|EPI_ISL_717945\|2020-1 | ···················· | ···················· | ····················· | ···················· | ···················· | ···················· |
| X | 4505 | hCoV-19/Brazil/RJ-00562/2020\|EPI_ISL_717946\|2020-1 | ···················· | ···················· | ····················· | ···················· | ···················· | ···················· |
| X | 4506 | hCoV-19/Brazil/RJ-00563/2020\|EPI_ISL_717947\|2020-1 | ···················· | ···················· | ····················· | ···················· | ···················· | ···················· |
| X | 4507 | hCoV-19/Brazil/RJ-00564/2020\|EPI_ISL_717948\|2020-1 | ···················· | ···················· | ····················· | ···················· | ···················· | ···················· |
| X | 4508 | hCoV-19/Brazil/RJ-00565/2020\|EPI_ISL_717949\|2020-1 | ···················· | ···················· | ····················· | ···················· | ···················· | ···················· |
| X | 4509 | hCoV-19/Brazil/RJ-00566/2020\|EPI_ISL_717950\|2020-1 | ···················· | ···················· | ····················· | ···················· | ···················· | ···················· |
| X | 4510 | hCoV-19/Brazil/RJ-00567/2020\|EPI_ISL_717951\|2020-1 | ···················· | ···················· | ····················· | ···················· | ···················· | ···················· |
| X | 4511 | hCoV-19/Brazil/RJ-00568/2020\|EPI_ISL_717952\|2020-1 | ···················· | ···················· | ····················· | ···················· | ···················· | ···················· |
| X | 4512 | hCoV-19/Brazil/RJ-00569/2020\|EPI_ISL_717953\|2020-1 | ···················· | ···················· | ····················· | ···················· | ···················· | ···················· |
| X | 4513 | hCoV-19/Brazil/RJ-00570/2020\|EPI_ISL_717954\|2020-1 | ···················· | ···················· | ····················· | ···················· | ···················· | ···················· |
| X | 4514 | hCoV-19/Brazil/RJ-00571/2020\|EPI_ISL_717955\|2020-1 | ···················· | ···················· | ····················· | ···················· | ···················· | ···················· |
| X | 4515 | hCoV-19/Brazil/RJ-00572/2020\|EPI_ISL_717956\|2020-1 | ···················· | ···················· | ····················· | ···················· | ···················· | ···················· |
| X | 4516 | hCoV-19/Brazil/RJ-00573/2020\|EPI_ISL_717957\|2020-1 | ···················· | ···················· | ····················· | ···················· | ···················· | ···················· |
| X | 4517 | hCoV-19/Brazil/RJ-00574/2020\|EPI_ISL_717958\|2020-0 | ···················· | ···················· | ····················· | ···················· | ···················· | ···················· |
| X | 4518 | hCoV-19/Brazil/RJ-00575/2020\|EPI_ISL_717959\|2020-0 | ···················· | ···················· | ····················· | ···················· | ···················· | ···················· |
| X | 4519 | hCoV-19/Brazil/RJ-00576/2020\|EPI_ISL_717960\|2020-1 | ···················· | ···················· | ····················· | ···················· | ···················· | ···················· |
| X | 4520 | hCoV-19/Brazil/RJ-00577/2020\|EPI_ISL_717961\|2020-1 | ···················· | ···················· | ····················· | ···················· | ···················· | ···················· |
| X | 4521 | hCoV-19/Brazil/RJ-00578/2020\|EPI_ISL_717962\|2020-0 | ···················· | ···················· | ····················· | ···················· | ···················· | ···················· |
| X | 4522 | hCoV-19/Brazil/RJ-00579/2020\|EPI_ISL_717963\|2020-0 | ···················· | ···················· | ····················· | ···················· | ···················· | ···················· |
| X | 4523 | hCoV-19/Brazil/RJ-00580/2020\|EPI_ISL_717964\|2020-0 | ···················· | ···················· | ····················· | ···················· | ···················· | ···················· |
| X | 4524 | hCoV-19/Russia/MOS-CRIE-13449451/2020\|EPI_ISL_7179 | ···················· | ···················· | ····················· | ···················· | ···················· | ···················· |
| X | 4525 | hCoV-19/Russia/MOS-CRIE-13455107/2020\|EPI_ISL_7179 | ···················· | ···················· | ····················· | ···················· | ···················· | ···················· |
| X | 4526 | hCoV-19/Russia/MOS-CRIE-13294236/2020\|EPI_ISL_7179 | ···················· | ···················· | ····················· | ···················· | ···················· | ···················· |
| X | 4527 | hCoV-19/Russia/MOS-CRIE-13297130/2020\|EPI_ISL_7179 | ···················· | ···················· | ····················· | ···················· | ···················· | ···················· |
| X | 4528 | hCoV-19/Russia/MOS-CRIE-13454908/2020\|EPI_ISL_7179 | ···················· | ···················· | ····················· | ···················· | ···················· | ···················· |
| X | 4529 | hCoV-19/Russia/MOS-CRIE-13379339/2020\|EPI_ISL_7179 | ···················· | ···················· | ····················· | ···················· | ···················· | ···················· |
| X | 4530 | hCoV-19/Russia/MOS-CRIE-13383171/2020\|EPI_ISL_7179 | ···················· | ···················· | ····················· | ···················· | ···················· | ···················· |
| X | 4531 | hCoV-19/Russia/MOS-CRIE-13312464/2020\|EPI_ISL_7179 | ···················· | ···················· | ····················· | ···················· | ···················· | ···················· |
| X | 4532 | hCoV-19/Russia/MOS-CRIE-13385510/2020\|EPI_ISL_7179 | ···················· | ···················· | ····················· | ···················· | ···················· | ···················· |
| X | 4533 | hCoV-19/Russia/MOS-CRIE-13386579/2020\|EPI_ISL_7179 | ···················· | ···················· | ·········T··········· | ···················· | ···················· | ···················· |
| X | 4534 | hCoV-19/cat/Greece/2K/2020\|EPI_ISL_717979\|2020-11- | ···················· | ···················· | ····················· | ···················· | ···················· | ···················· |
| X | 4535 | hCoV-19/Belgium/UGent-251/2020\|EPI_ISL_717991\|2020 | ···················· | ···················· | ····················· | ···················· | ···················· | ···················· |
| X | 4536 | hCoV-19/Belgium/UGent-252/2020\|EPI_ISL_717992\|2020 | ···················· | ···················· | ····················· | ···················· | ······T············· | ···················· |
| X | 4537 | hCoV-19/Belgium/UGent-253/2020\|EPI_ISL_717993\|2020 | ···················· | ···················· | ····················· | ···················· | ···················· | ···················· |
| X | 4538 | hCoV-19/Belgium/UGent-254/2020\|EPI_ISL_717994\|2020 | ···················· | ···················· | ····················· | ···················· | ···················· | ···················· |
| X | 4540 | hCoV-19/Belgium/UGent-256/2020\|EPI_ISL_717996\|2020 | ···················· | ···················· | ····················· | ···················· | ···················· | ···················· |
| X | 4541 | hCoV-19/Belgium/UGent-257/2020\|EPI_ISL_717997\|2020 | ···················· | ···················· | ····················· | ···················· | ···················· | ···················· |
| X | 4542 | hCoV-19/Belgium/UGent-258/2020\|EPI_ISL_717998\|2020 | ···················· | ···················· | ····················· | ···················· | ···················· | ···················· |
| X | 4543 | hCoV-19/Belgium/UGent-259/2020\|EPI_ISL_717999\|2020 | ···················· | ···················· | ····················· | ···················· | ···················· | ···················· |
| X | 4544 | hCoV-19/England/QEUH-BA3990/2020\|EPI_ISL_721129\|20 | ···················· | ···················· | ····················· | ···················· | ···················· | ···················· |
| X | 4545 | hCoV-19/England/QEUH-BA3295/2020\|EPI_ISL_721130\|20 | ···················· | ···················· | ····················· | ···················· | ···················· | ···················· |
| X | 4546 | hCoV-19/England/QEUH-B9E1DC/2020\|EPI_ISL_721131\|20 | ···················· | ···················· | ····················· | ···················· | ···················· | ···················· |
| X | 4547 | hCoV-19/England/QEUH-BA32B3/2020\|EPI_ISL_721161\|20 | ···················· | ···················· | ····················· | ···················· | ···················· | ···················· |
| X | 4548 | hCoV-19/England/QEUH-B9F654/2020\|EPI_ISL_721162\|20 | ···················· | ···················· | ····················· | ···················· | ···················· | ···················· |
| X | 4549 | hCoV-19/England/QEUH-BA3D2B/2020\|EPI_ISL_721163\|20 | ···················· | ···················· | ····················· | ···················· | ···················· | ···················· |
| X | 4550 | hCoV-19/England/QEUH-B9FDB9/2020\|EPI_ISL_721164\|20 | ···················· | ···················· | ····················· | ···················· | ···················· | ···················· |
| X | 4551 | hCoV-19/England/QEUH-B9F91F/2020\|EPI_ISL_721165\|20 | ···················· | ···················· | ····················· | ···················· | ···················· | ···················· |
| X | 4552 | hCoV-19/England/QEUH-BA41C4/2020\|EPI_ISL_721166\|20 | ···················· | ···················· | ····················· | ···················· | ···················· | ···················· |
| X | 4553 | hCoV-19/England/QEUH-B9F8B8/2020\|EPI_ISL_721168\|20 | ···················· | ···················· | ····················· | ···················· | ···················· | ···················· |
| X | 4554 | hCoV-19/England/QEUH-B9E127/2020\|EPI_ISL_721169\|20 | ···················· | ···················· | ····················· | ···················· | ···················· | ···················· |
| X | 4555 | hCoV-19/England/QEUH-BA3329/2020\|EPI_ISL_721171\|20 | ···················· | ···················· | ····················· | ···················· | ···················· | ···················· |
| X | 4556 | hCoV-19/England/QEUH-B9FA1C/2020\|EPI_ISL_721172\|20 | ···················· | ···················· | ····················· | ···················· | ···················· | ···················· |
| X | 4557 | hCoV-19/England/QEUH-B9F9A6/2020\|EPI_ISL_721174\|20 | ···················· | ···················· | ····················· | ···················· | ···················· | ···················· |
| X | 4558 | hCoV-19/England/QEUH-BA363F/2020\|EPI_ISL_721175\|20 | ···················· | ···················· | ····················· | ···················· | ···················· | ···················· |
| X | 4559 | hCoV-19/England/QEUH-B9FBDD/2020\|EPI_ISL_721176\|20 | ···················· | ···················· | ····················· | ···················· | ···················· | ···················· |
| X | 4560 | hCoV-19/England/QEUH-B9F33F/2020\|EPI_ISL_721177\|20 | ···················· | ···················· | ····················· | ···················· | ···················· | ···················· |
| X | 4561 | hCoV-19/England/QEUH-BA4285/2020\|EPI_ISL_721178\|20 | ···················· | ···················· | ····················· | ···················· | ···················· | ···················· |
| X | 4562 | hCoV-19/England/QEUH-BA4188/2020\|EPI_ISL_721179\|20 | ···················· | ···················· | ····················· | ···················· | ···················· | ···················· |
| X | 4563 | hCoV-19/England/QEUH-BA4267/2020\|EPI_ISL_721180\|20 | ···················· | ···················· | ····················· | ···················· | ···················· | ···················· |
| X | 4564 | hCoV-19/England/QEUH-BA4276/2020\|EPI_ISL_721181\|20 | ···················· | ···················· | ····················· | ···················· | ···················· | ···················· |
| X | 4565 | hCoV-19/England/QEUH-BA41F1/2020\|EPI_ISL_721182\|20 | ···················· | ···················· | ····················· | ···················· | ···················· | ··A················· |
| X | 4566 | hCoV-19/England/QEUH-B9F5A2/2020\|EPI_ISL_721183\|20 | ···················· | ···················· | ····················· | ···················· | ···················· | ···················· |
| X | 4567 | hCoV-19/England/QEUH-BA416A/2020\|EPI_ISL_721184\|20 | ···················· | ···················· | ····················· | ···················· | ···················· | ···················· |
| X | 4568 | hCoV-19/England/ALDP-BC76BD/2020\|EPI_ISL_721185\|20 | ···················· | ···················· | ····················· | ···················· | ···················· | ···················· |
| X | 4569 | hCoV-19/England/ALDP-BC66BE/2020\|EPI_ISL_721187\|20 | ···················· | ···················· | ····················· | ···················· | ···················· | ···················· |
| X | 4570 | hCoV-19/England/ALDP-BC831F/2020\|EPI_ISL_721190\|20 | ···················· | ···················· | ····················· | ···················· | ···················· | ···················· |
| X | 4571 | hCoV-19/England/ALDP-BC74D1/2020\|EPI_ISL_721192\|20 | ···················· | ···················· | ····················· | ···················· | ······T············· | ···················· |
| X | 4572 | hCoV-19/England/ALDP-BC685E/2020\|EPI_ISL_721194\|20 | ···················· | ···················· | ····················· | ···················· | ···················· | ···················· |
| X | 4573 | hCoV-19/England/ALDP-BC6250/2020\|EPI_ISL_721195\|20 | ···················· | ···················· | ····················· | ···················· | ···················· | ···················· |
| X | 4574 | hCoV-19/England/ALDP-BC86E9/2020\|EPI_ISL_721196\|20 | ···················· | ···················· | ····················· | ···················· | ···················· | ···················· |
| X | 4575 | hCoV-19/England/ALDP-BC73D4/2020\|EPI_ISL_721197\|20 | ···················· | ···················· | ····················· | ···················· | ···················· | ···················· |
| X | 4576 | hCoV-19/England/ALDP-BC83C4/2020\|EPI_ISL_721198\|20 | ···················· | ···················· | ····················· | ···················· | ···················· | ···················· |
| X | 4577 | hCoV-19/England/ALDP-BC6609/2020\|EPI_ISL_721199\|20 | ···················· | ···················· | ····················· | ···················· | ···················· | ···················· |
| X | 4578 | hCoV-19/Netherlands/UT-EMC-228/2020\|EPI_ISL_722460 | ···················· | ···················· | ····················· | ···················· | ···················· | ···················· |

| | | | | | | | |
|---|---|---|---|---|---|---|---|
| X | 4579 hCoV-19/Netherlands/UT-EMC-222/2020\|EPI_ISL_722461 | ···················· | ···················· | ···················· | ···················· | ···················· | ···················· |
| X | 4580 hCoV-19/Netherlands/UT-EMC-223/2020\|EPI_ISL_722462 | ···················· | ···················· | ···················· | ···················· | ···················· | ···················· |
| X | 4581 hCoV-19/Netherlands/ZH-EMC-938/2020\|EPI_ISL_722490 | ···················· | ···················· | ···················· | ···················· | ···················· | ···················· |
| X | 4582 hCoV-19/Netherlands/ZH-EMC-939/2020\|EPI_ISL_722491 | ···················· | ···················· | ···················· | ···················· | ···················· | ···················· |
| X | 4583 hCoV-19/Netherlands/NB-EMC-370/2020\|EPI_ISL_722492 | ···················· | ···················· | ···················· | ···················· | ···················· | ···················· |
| X | 4584 hCoV-19/Netherlands/ZH-EMC-1067/2020\|EPI_ISL_72249 | ···················· | ···················· | ···················· | ···················· | ···················· | ···················· |
| X | 4585 hCoV-19/Netherlands/ZH-EMC-943/2020\|EPI_ISL_722494 | ···················· | ···················· | ···················· | ···················· | ···················· | ···················· |
| X | 4586 hCoV-19/Netherlands/ZH-EMC-971/2020\|EPI_ISL_722495 | ···················· | ···················· | ···················· | ···················· | ···················· | ···················· |
| X | 4587 hCoV-19/Netherlands/NB-EMC-324/2020\|EPI_ISL_722496 | ···················· | ···················· | ···················· | ···················· | ···················· | ···················· |
| X | 4588 hCoV-19/Netherlands/GE-EMC-355/2020\|EPI_ISL_722497 | ···················· | ···················· | ···················· | ···················· | ···················· | ···················· |
| X | 4589 hCoV-19/Netherlands/GE-EMC-379/2020\|EPI_ISL_722498 | ···················· | ···················· | ···················· | ···················· | ···················· | ···················· |
| X | 4590 hCoV-19/Netherlands/ZH-EMC-940/2020\|EPI_ISL_722499 | ···················· | ···················· | ···················· | ···················· | ···················· | ···················· |
| X | 4591 hCoV-19/England/ALDP-B76996/2020\|EPI_ISL_723790\|20 | ···················· | ···················· | ···················· | ···················· | ···················· | ···················· |
| X | 4592 hCoV-19/England/ALDP-B769A5/2020\|EPI_ISL_723791\|20 | ···················· | ···················· | ···················· | ···················· | ···················· | ···················· |
| X | 4593 hCoV-19/England/ALDP-B769B4/2020\|EPI_ISL_723792\|20 | ···················· | ···················· | ···················· | ···················· | ···················· | ···················· |
| X | 4594 hCoV-19/England/ALDP-B769C3/2020\|EPI_ISL_723793\|20 | ···················· | ···················· | ···················· | ···················· | ···················· | ···················· |
| X | 4595 hCoV-19/England/ALDP-B769F0/2020\|EPI_ISL_723796\|20 | ···················· | ···················· | ···················· | ···················· | ···················· | ···················· |
| X | 4596 hCoV-19/England/ALDP-B76A66/2020\|EPI_ISL_723800\|20 | ···················· | ···················· | ···················· | ···················· | ···················· | ···················· |
| X | 4597 hCoV-19/England/ALDP-B76A75/2020\|EPI_ISL_723801\|20 | ···················· | ···················· | ···················· | ···················· | ···················· | ···················· |
| X | 4598 hCoV-19/England/ALDP-B76A93/2020\|EPI_ISL_723802\|20 | ···················· | ···················· | ···················· | ···················· | ······T············· | ···················· |
| X | 4599 hCoV-19/England/ALDP-B76AC0/2020\|EPI_ISL_723803\|20 | ···················· | ···················· | ···················· | ···················· | ···················· | ···················· |
| X | 4600 hCoV-19/England/ALDP-B76CE8/2020\|EPI_ISL_723804\|20 | ···················· | ···················· | ···················· | ···················· | ···················· | ···················· |
| X | 4601 hCoV-19/England/ALDP-B76CF7/2020\|EPI_ISL_723805\|20 | ···················· | ···················· | ···················· | ···················· | ···················· | ···················· |
| X | 4602 hCoV-19/England/ALDP-B76D30/2020\|EPI_ISL_723806\|20 | ···················· | ···················· | ···················· | ···················· | ···················· | ···················· |
| X | 4603 hCoV-19/England/ALDP-B76D4F/2020\|EPI_ISL_723807\|20 | ···················· | ···················· | ···················· | ···················· | ···················· | ···················· |
| X | 4604 hCoV-19/England/ALDP-B76D5E/2020\|EPI_ISL_723808\|20 | ···················· | ·A·················· | ···················· | ···················· | ···················· | ··A················· |
| X | 4605 hCoV-19/England/ALDP-B76D6D/2020\|EPI_ISL_723809\|20 | ···················· | ···················· | ···················· | ···················· | ···················· | ···················· |
| X | 4606 hCoV-19/England/ALDP-B76D7C/2020\|EPI_ISL_723810\|20 | ···················· | ···················· | ···················· | ···················· | ···················· | ···················· |
| X | 4607 hCoV-19/England/ALDP-B76DA9/2020\|EPI_ISL_723811\|20 | ···················· | ···················· | ···················· | ···················· | ···················· | ···················· |
| X | 4608 hCoV-19/England/ALDP-B76DB8/2020\|EPI_ISL_723812\|20 | ···················· | ···················· | ···················· | ···················· | ···················· | ···················· |
| X | 4609 hCoV-19/Italy/SAR-ASSL/2020\|EPI_ISL_710503\|2020-10 | ···················· | ···················· | ···················· | ···················· | ···················· | ···················· |
| X | 4610 hCoV-19/Australia/NSW3666/2020\|EPI_ISL_710513\|2020 | ···················· | ···················· | ···················· | ···················· | ···················· | ···················· |
| X | 4611 hCoV-19/England/204980392/2020\|EPI_ISL_710515\|2020 | ···················· | ···················· | ···················· | ···················· | ···················· | ···················· |
| X | 4612 hCoV-19/England/204980417/2020\|EPI_ISL_710516\|2020 | ···················· | ···················· | ···················· | ···················· | ···················· | ···················· |
| X | 4613 hCoV-19/England/204990417/2020\|EPI_ISL_710517\|2020 | ···················· | ···················· | ···················· | ···················· | ······T············· | ···················· |
| X | 4614 hCoV-19/England/204990708/2020\|EPI_ISL_710518\|2020 | ···················· | ···················· | ···················· | ···················· | ······T············· | ···················· |
| X | 4615 hCoV-19/England/205020882/2020\|EPI_ISL_710519\|2020 | ···················· | ···················· | ···················· | ···················· | ···················· | ···················· |
| X | 4616 hCoV-19/England/205020883/2020\|EPI_ISL_710520\|2020 | ···················· | ···················· | ···················· | ···················· | ···················· | ···················· |
| X | 4617 hCoV-19/England/205020884/2020\|EPI_ISL_710521\|2020 | ···················· | ···················· | ···················· | ···················· | ···················· | ···················· |
| X | 4618 hCoV-19/England/205041756/2020\|EPI_ISL_710525\|2020 | ···················· | ···················· | ···················· | ···················· | ···················· | ···················· |
| X | 4619 hCoV-19/England/205041759/2020\|EPI_ISL_710526\|2020 | ···················· | ···················· | ···················· | ···················· | ···················· | ···················· |
| X | 4620 hCoV-19/England/205041761/2020\|EPI_ISL_710527\|2020 | ···················· | ···················· | ···················· | ···················· | ···················· | ···················· |
| X | 4621 hCoV-19/England/205041767/2020\|EPI_ISL_710529\|2020 | ···················· | ···················· | ···················· | ···················· | ···················· | ···················· |
| X | 4622 hCoV-19/Tunisia/ADAGE-12622/2020\|EPI_ISL_710532\|20 | ···················· | ···················· | ···················· | ···················· | ···················· | ···················· |
| X | 4623 hCoV-19/Tunisia/ADAGE-17873/2020\|EPI_ISL_710534\|20 | ···················· | ···················· | ···················· | ···················· | ···················· | ···················· |
| X | 4624 hCoV-19/Gibraltar/205000629/2020\|EPI_ISL_710535\|20 | ···················· | ···················· | ···················· | ···················· | ···················· | ···················· |
| X | 4625 hCoV-19/Tunisia/ADAGE-17872/2020\|EPI_ISL_710537\|20 | ···················· | ···················· | ···················· | ···················· | ··········T········· | ···················· |
| X | 4626 hCoV-19/Tunisia/ADAGE-18212/2020\|EPI_ISL_710540\|20 | ···················· | ···················· | ···················· | ···················· | ···················· | ···················· |
| X | 4627 hCoV-19/Tunisia/ADAGE-18245/2020\|EPI_ISL_710541\|20 | ···················· | ···················· | ···················· | ···················· | ···················· | ···················· |
| X | 4628 hCoV-19/Italy/LAZ-INMI-92/2020\|EPI_ISL_710542\|2020 | ···················· | ···················· | ···················· | ···················· | ···················· | ···················· |
| X | 4629 hCoV-19/Italy/LAZ-INMI-93/2020\|EPI_ISL_710543\|2020 | ···················· | ···················· | ···················· | ···················· | ···················· | ···················· |
| X | 4631 hCoV-19/Italy/LAZ-INMI-95/2020\|EPI_ISL_710545\|2020 | ···················· | ···················· | ···················· | ···················· | ···················· | ···················· |
| X | 4632 hCoV-19/Italy/LAZ-INMI-96/2020\|EPI_ISL_710546\|2020 | ···················· | ···················· | ···················· | ···················· | ···················· | ···················· |
| X | 4633 hCoV-19/Croatia/Zagreb-27/2020\|EPI_ISL_710547\|2020 | ···················· | ···················· | ···················· | ···················· | ···················· | ···················· |
| X | 4634 hCoV-19/Croatia/Zagreb-15/2020\|EPI_ISL_710548\|2020 | ···················· | ···················· | ···················· | ···················· | ···················· | ···················· |
| X | 4635 hCoV-19/Croatia/Zagreb-12/2020\|EPI_ISL_710549\|2020 | ···················· | ···················· | ···················· | ···················· | ···················· | ···················· |
| X | 4636 hCoV-19/Croatia/Zagreb-29/2020\|EPI_ISL_710554\|2020 | ···················· | ···················· | ···················· | ···················· | ···················· | ···················· |
| X | 4637 hCoV-19/Croatia/Zagreb-14/2020\|EPI_ISL_710555\|2020 | ···················· | ···················· | ···················· | ···················· | ···················· | ···················· |
| X | 4638 hCoV-19/Croatia/Zagreb-4/2020\|EPI_ISL_710556\|2020- | ···················· | ···················· | ···················· | ···················· | ···················· | ···················· |
| X | 4639 hCoV-19/Croatia/Zagreb-7/2020\|EPI_ISL_710557\|2020- | ···················· | ···················· | ···················· | ···················· | ···················· | ···················· |
| X | 4640 hCoV-19/Croatia/Zagreb-11/2020\|EPI_ISL_710569\|2020 | ···················· | ···················· | ···················· | ···················· | ···················· | ···················· |
| X | 4641 hCoV-19/Croatia/Zagreb-22/2020\|EPI_ISL_710570\|2020 | ···················· | ···················· | ···················· | ···················· | ···················· | ···················· |
| X | 4642 hCoV-19/Croatia/Zagreb-5/2020\|EPI_ISL_710571\|2020- | ···················· | ···················· | ···················· | ···················· | ···················· | ···················· |
| X | 4643 hCoV-19/Croatia/Zagreb-8/2020\|EPI_ISL_710572\|2020- | ···················· | ···················· | ···················· | ···················· | ···················· | ···················· |
| X | 4644 hCoV-19/Croatia/Zagreb-26/2020\|EPI_ISL_710573\|2020 | ···················· | ···················· | ···················· | ···················· | ···················· | ···················· |
| X | 4645 hCoV-19/Croatia/Zagreb-1/2020\|EPI_ISL_710574\|2020- | ···················· | ···················· | ···················· | ···················· | ···················· | ···················· |
| X | 4646 hCoV-19/Tunisia/ADAGE-17786/2020\|EPI_ISL_710575\|20 | ···················· | ···················· | ···················· | ···················· | ···················· | ···················· |
| X | 4647 hCoV-19/Sweden/20-53570/2020\|EPI_ISL_710576\|2020-1 | ···················· | ···················· | ···················· | ···················· | ···················· | ···················· |
| X | 4648 hCoV-19/Sweden/20-24575/2020\|EPI_ISL_710577\|2020-1 | ···················· | ···················· | ···················· | ···················· | ···················· | ···················· |
| X | 4649 hCoV-19/Sweden/20-24735/2020\|EPI_ISL_710578\|2020-1 | ···················· | ···················· | ···················· | ···················· | ···················· | ···················· |
| X | 4650 hCoV-19/Sweden/20-23090/2020\|EPI_ISL_710579\|2020-0 | ···················· | ···················· | ···················· | ···················· | ···················· | ···················· |
| X | 4651 hCoV-19/Sweden/20-24262/2020\|EPI_ISL_710580\|2020-0 | ···················· | ···················· | ···················· | ···················· | ···················· | ···················· |
| X | 4652 hCoV-19/Sweden/20-53564/2020\|EPI_ISL_710581\|2020-1 | ···················· | ···················· | ···················· | ···················· | ···················· | ···················· |
| X | 4653 hCoV-19/Sweden/20-53565/2020\|EPI_ISL_710582\|2020-1 | ···················· | ···················· | ···················· | ···················· | ···················· | ···················· |
| X | 4654 hCoV-19/Sweden/20-53566/2020\|EPI_ISL_710583\|2020-1 | ···················· | ···················· | ···················· | ···················· | ···················· | ···················· |
| X | 4655 hCoV-19/Sweden/20-08532/2020\|EPI_ISL_710584\|2020-0 | ···················· | ···················· | ···················· | ···················· | ···················· | ···················· |
| X | 4656 hCoV-19/Sweden/20-24683/2020\|EPI_ISL_710585\|2020-1 | ···················· | ···················· | ···················· | ···················· | ······T············· | ···················· |
| X | 4657 hCoV-19/Sweden/20-24685/2020\|EPI_ISL_710586\|2020-1 | ···················· | ···················· | ···················· | ···················· | ······T············· | ···················· |
| X | 4658 hCoV-19/Sweden/20-52399/2020\|EPI_ISL_710587\|2020-0 | ···················· | ···················· | ···················· | ···················· | ···················· | ···················· |
| X | 4659 hCoV-19/Sweden/20-53552/2020\|EPI_ISL_710588\|2020-1 | ···················· | ···················· | ···················· | ···················· | ···················· | ···················· |

| | | | | | | | |
|---|---|---|---|---|---|---|---|
| X | 4660 hCoV-19/Sweden/20-53470/2020\|EPI_ISL_710589\|2020-1 | ···················· | ···················· | ···················· | ···················· | ···················· | ···················· |
| X | 4661 hCoV-19/Denmark/DCGC-14502/2020\|EPI_ISL_711801\|202 | ···················· | ···················· | ···················· | ···················· | ···················· | ···················· |
| X | 4662 hCoV-19/Denmark/DCGC-14504/2020\|EPI_ISL_711802\|202 | ···················· | ···················· | ···················· | ···················· | ···················· | ···················· |
| X | 4663 hCoV-19/Denmark/DCGC-14505/2020\|EPI_ISL_711803\|202 | ···················· | ···················· | ···················· | ···················· | ···················· | ···················· |
| X | 4664 hCoV-19/Denmark/DCGC-14506/2020\|EPI_ISL_711804\|202 | ···················· | ···················· | ···················· | ···················· | ···················· | ···················· |
| X | 4665 hCoV-19/Denmark/DCGC-14507/2020\|EPI_ISL_711805\|202 | ···················· | ···················· | ···················· | ···················· | ···················· | ···················· |
| X | 4666 hCoV-19/Denmark/DCGC-14508/2020\|EPI_ISL_711806\|202 | ···················· | ···················· | ···················· | ···················· | ···················· | ···················· |
| X | 4667 hCoV-19/Denmark/DCGC-14509/2020\|EPI_ISL_711807\|202 | ···················· | ···················· | ···················· | ···················· | ···················· | ···················· |
| X | 4668 hCoV-19/Denmark/DCGC-14511/2020\|EPI_ISL_711808\|202 | ···················· | ···················· | ···················· | ···················· | ···················· | ···················· |
| X | 4669 hCoV-19/Denmark/DCGC-14512/2020\|EPI_ISL_711809\|202 | ···················· | ···················· | ···················· | ···················· | ···················· | ···················· |
| X | 4670 hCoV-19/Denmark/DCGC-14513/2020\|EPI_ISL_711810\|202 | ···················· | ···················· | ···················· | ···················· | ···················· | ···················· |
| X | 4671 hCoV-19/Denmark/DCGC-14514/2020\|EPI_ISL_711811\|202 | ···················· | ·A·················· | ···················· | ···················· | ···················· | ···················· |
| X | 4672 hCoV-19/Denmark/DCGC-14552/2020\|EPI_ISL_711834\|202 | ···················· | ···················· | ···················· | ···················· | ···················· | ···················· |
| X | 4673 hCoV-19/Denmark/DCGC-14553/2020\|EPI_ISL_711835\|202 | ···················· | ···················· | ···················· | ···················· | ···················· | ···················· |
| X | 4674 hCoV-19/Denmark/DCGC-14554/2020\|EPI_ISL_711836\|202 | ···················· | ···················· | ···················· | ···················· | ···················· | ···················· |
| X | 4675 hCoV-19/Denmark/DCGC-14555/2020\|EPI_ISL_711837\|202 | ···················· | ···················· | ···················· | ···················· | ···················· | ···················· |
| X | 4676 hCoV-19/Denmark/DCGC-14557/2020\|EPI_ISL_711838\|202 | ···················· | ···················· | ···················· | ···················· | ···················· | ···················· |
| X | 4677 hCoV-19/Denmark/DCGC-14558/2020\|EPI_ISL_711839\|202 | ···················· | ···················· | ···················· | ···················· | ···················· | ···················· |
| X | 4678 hCoV-19/Denmark/DCGC-14560/2020\|EPI_ISL_711840\|202 | ···················· | ···················· | ···················· | ···················· | ···················· | ···················· |
| X | 4679 hCoV-19/Denmark/DCGC-14566/2020\|EPI_ISL_711841\|202 | ···················· | ·A·················· | ···················· | ···················· | ···················· | ···················· |
| X | 4680 hCoV-19/Denmark/DCGC-14567/2020\|EPI_ISL_711842\|202 | ···················· | ···················· | ···················· | ···················· | ···················· | ···················· |
| X | 4681 hCoV-19/Denmark/DCGC-14569/2020\|EPI_ISL_711843\|202 | ···················· | ···················· | ···················· | ···················· | ···················· | ···················· |
| X | 4682 hCoV-19/Denmark/DCGC-14570/2020\|EPI_ISL_711844\|202 | ···················· | ···················· | ···················· | ···················· | ···················· | ···················· |
| X | 4683 hCoV-19/Denmark/DCGC-14571/2020\|EPI_ISL_711845\|202 | ···················· | ···················· | ···················· | ···················· | ···················· | ···················· |
| X | 4684 hCoV-19/Denmark/DCGC-14572/2020\|EPI_ISL_711846\|202 | ···················· | ···················· | ···················· | ···················· | ···················· | ···················· |
| X | 4685 hCoV-19/Denmark/DCGC-14573/2020\|EPI_ISL_711847\|202 | ···················· | ···················· | ···················· | ···················· | ···················· | ···················· |
| X | 4686 hCoV-19/Denmark/DCGC-14575/2020\|EPI_ISL_711848\|202 | ···················· | ···················· | ···················· | ···················· | ···················· | ···················· |
| X | 4687 hCoV-19/Denmark/DCGC-14576/2020\|EPI_ISL_711849\|202 | ···················· | ···················· | ···················· | ···················· | ···················· | ···················· |
| X | 4688 hCoV-19/Denmark/DCGC-14577/2020\|EPI_ISL_711850\|202 | ···················· | ···················· | ···················· | ···················· | ···················· | ···················· |
| X | 4689 hCoV-19/Denmark/DCGC-14580/2020\|EPI_ISL_711851\|202 | ···················· | ···················· | ···················· | ···················· | ···················· | ···················· |
| X | 4690 hCoV-19/Denmark/DCGC-14581/2020\|EPI_ISL_711852\|202 | ···················· | ···················· | ···················· | ···················· | ···················· | ···················· |
| X | 4691 hCoV-19/Denmark/DCGC-14582/2020\|EPI_ISL_711853\|202 | ···················· | ···················· | ···················· | ···················· | ···················· | ···················· |
| X | 4692 hCoV-19/Denmark/DCGC-14583/2020\|EPI_ISL_711854\|202 | ···················· | ···················· | ···················· | ···················· | ···················· | ···················· |
| X | 4693 hCoV-19/Denmark/DCGC-14584/2020\|EPI_ISL_711855\|202 | ···················· | ···················· | ···················· | ···················· | ···················· | ···················· |
| X | 4694 hCoV-19/Denmark/DCGC-14585/2020\|EPI_ISL_711856\|202 | ···················· | ···················· | ···················· | ···················· | ···················· | ···················· |
| X | 4695 hCoV-19/Denmark/DCGC-14586/2020\|EPI_ISL_711857\|202 | ···················· | ···················· | ···················· | ···················· | ···················· | ···················· |
| X | 4696 hCoV-19/Denmark/DCGC-14587/2020\|EPI_ISL_711858\|202 | ···················· | ···················· | ···················· | ···················· | ···················· | ···················· |
| X | 4697 hCoV-19/Denmark/DCGC-14588/2020\|EPI_ISL_711859\|202 | ···················· | ···················· | ···················· | ···················· | ···················· | ···················· |
| X | 4698 hCoV-19/Denmark/DCGC-14589/2020\|EPI_ISL_711860\|202 | ···················· | ·A·················· | ···················· | ···················· | ···················· | ··A················· |
| X | 4699 hCoV-19/Denmark/DCGC-14591/2020\|EPI_ISL_711861\|202 | ···················· | ···················· | ···················· | ···················· | ···················· | ···················· |
| X | 4700 hCoV-19/Denmark/DCGC-14592/2020\|EPI_ISL_711862\|202 | ···················· | ···················· | ···················· | ···················· | ···················· | ···················· |
| X | 4701 hCoV-19/Denmark/DCGC-14593/2020\|EPI_ISL_711863\|202 | ···················· | ···················· | ···················· | ···················· | ···················· | ···················· |
| X | 4702 hCoV-19/Denmark/DCGC-14595/2020\|EPI_ISL_711864\|202 | ···················· | ···················· | ···················· | ···················· | ···················· | ···················· |
| X | 4703 hCoV-19/Denmark/DCGC-14597/2020\|EPI_ISL_711865\|202 | ···················· | ···················· | ···················· | ···················· | ···················· | ···················· |
| X | 4704 hCoV-19/Denmark/DCGC-14598/2020\|EPI_ISL_711866\|202 | ···················· | ···················· | ···················· | ···················· | ···················· | ···················· |
| X | 4705 hCoV-19/Denmark/DCGC-14602/2020\|EPI_ISL_711867\|202 | ···················· | ···················· | ···················· | ···················· | ···················· | ···················· |
| X | 4706 hCoV-19/Denmark/DCGC-14604/2020\|EPI_ISL_711868\|202 | ···················· | ···················· | ···················· | ···················· | ···················· | ···················· |
| X | 4707 hCoV-19/Denmark/DCGC-14607/2020\|EPI_ISL_711869\|202 | ···················· | ···················· | ···················· | ···················· | ···················· | ···················· |
| X | 4708 hCoV-19/Denmark/DCGC-14609/2020\|EPI_ISL_711870\|202 | ···················· | ···················· | ···················· | ···················· | ···················· | ···················· |
| X | 4709 hCoV-19/Denmark/DCGC-14612/2020\|EPI_ISL_711871\|202 | ···················· | ···················· | ···················· | ···················· | ···················· | ···················· |
| X | 4710 hCoV-19/Denmark/DCGC-14613/2020\|EPI_ISL_711872\|202 | ···················· | ···················· | ···················· | ···················· | ···················· | ···················· |
| X | 4711 hCoV-19/Denmark/DCGC-14618/2020\|EPI_ISL_711873\|202 | ···················· | ···················· | ···················· | ···················· | ···················· | ···················· |
| X | 4712 hCoV-19/Denmark/DCGC-14621/2020\|EPI_ISL_711874\|202 | ···················· | ···················· | ···················· | ···················· | ···················· | ···················· |
| X | 4713 hCoV-19/Denmark/DCGC-14622/2020\|EPI_ISL_711875\|202 | ···················· | ···················· | ···················· | ···················· | ···················· | ···················· |
| X | 4714 hCoV-19/Denmark/DCGC-14623/2020\|EPI_ISL_711876\|202 | ···················· | ···················· | ···················· | ···················· | ···················· | ···················· |
| X | 4715 hCoV-19/Denmark/DCGC-14627/2020\|EPI_ISL_711877\|202 | ···················· | ···················· | ···················· | ···················· | ···················· | ···················· |
| X | 4716 hCoV-19/Denmark/DCGC-14628/2020\|EPI_ISL_711878\|202 | ···················· | ···················· | ···················· | ···················· | ···················· | ···················· |
| X | 4717 hCoV-19/Denmark/DCGC-14629/2020\|EPI_ISL_711879\|202 | ···················· | ···················· | ···················· | ···················· | ···················· | ···················· |
| X | 4718 hCoV-19/Denmark/DCGC-14635/2020\|EPI_ISL_711883\|202 | ···················· | ···················· | ···················· | ···················· | ···················· | ···················· |
| X | 4719 hCoV-19/Denmark/DCGC-14637/2020\|EPI_ISL_711885\|202 | ···················· | ···················· | ···················· | ···················· | ···················· | ···················· |
| X | 4720 hCoV-19/Denmark/DCGC-14639/2020\|EPI_ISL_711886\|202 | ···················· | ···················· | ···················· | ···················· | ···················· | ···················· |
| X | 4721 hCoV-19/Denmark/DCGC-14642/2020\|EPI_ISL_711887\|202 | ···················· | ···················· | ···················· | ···················· | ···················· | ···················· |
| X | 4722 hCoV-19/Denmark/DCGC-14643/2020\|EPI_ISL_711888\|202 | ···················· | ·A·················· | ···················· | ···················· | ···················· | ···················· |
| X | 4723 hCoV-19/Denmark/DCGC-15867/2020\|EPI_ISL_714073\|202 | ···················· | ···················· | ···················· | ···················· | ···················· | ···················· |
| X | 4724 hCoV-19/Denmark/DCGC-15868/2020\|EPI_ISL_714074\|202 | ···················· | ···················· | ···················· | ···················· | ···················· | ···················· |
| X | 4725 hCoV-19/Denmark/DCGC-15869/2020\|EPI_ISL_714075\|202 | ···················· | ···················· | ···················· | ···················· | ···················· | ···················· |
| X | 4726 hCoV-19/Denmark/DCGC-15871/2020\|EPI_ISL_714076\|202 | ···················· | ···················· | ···················· | ···················· | ···················· | ···················· |
| X | 4727 hCoV-19/Denmark/DCGC-15872/2020\|EPI_ISL_714077\|202 | ···················· | ···················· | ···················· | ···················· | ···················· | ···················· |
| X | 4728 hCoV-19/Denmark/DCGC-15878/2020\|EPI_ISL_714080\|202 | ···················· | ···················· | ···················· | ···················· | ···················· | ···················· |
| X | 4729 hCoV-19/Denmark/DCGC-15880/2020\|EPI_ISL_714081\|202 | ···················· | ···················· | ···················· | ···················· | ···················· | ···················· |
| X | 4730 hCoV-19/Denmark/DCGC-15884/2020\|EPI_ISL_714082\|202 | ···················· | ···················· | ···················· | ···················· | ···················· | ···················· |
| X | 4731 hCoV-19/Denmark/DCGC-15885/2020\|EPI_ISL_714083\|202 | ···················· | ···················· | ···················· | ···················· | ···················· | ···················· |
| X | 4732 hCoV-19/Denmark/DCGC-15887/2020\|EPI_ISL_714084\|202 | ···················· | ···················· | ···················· | ···················· | ···················· | ···················· |
| X | 4733 hCoV-19/Denmark/DCGC-15888/2020\|EPI_ISL_714085\|202 | ···················· | ·A·················· | ···················· | ···················· | ···················· | ··A················· |
| X | 4734 hCoV-19/Denmark/DCGC-15889/2020\|EPI_ISL_714086\|202 | ···················· | ···················· | ···················· | ···················· | ···················· | ···················· |
| X | 4735 hCoV-19/Denmark/DCGC-15890/2020\|EPI_ISL_714087\|202 | ···················· | ···················· | ···················· | ···················· | ···················· | ···················· |
| X | 4736 hCoV-19/Denmark/DCGC-15893/2020\|EPI_ISL_714088\|202 | ···················· | ···················· | ···················· | ···················· | ···················· | ···················· |
| X | 4737 hCoV-19/Denmark/DCGC-15894/2020\|EPI_ISL_714089\|202 | ···················· | ···················· | ···················· | ···················· | ···················· | ···················· |
| X | 4738 hCoV-19/England/ALDP-BC7398/2020\|EPI_ISL_721200\|20 | ···················· | ···················· | ···················· | ···················· | ···················· | ···················· |
| X | 4739 hCoV-19/England/ALDP-BC786C/2020\|EPI_ISL_721201\|20 | ···················· | ···················· | ···················· | ···················· | ···················· | ···················· |

| | | | | | | | | |
|---|---|---|---|---|---|---|---|---|
| X | 4740 | hCoV-19/England/ALDP-BC6681/2020\|EPI_ISL_721202\|20 | ···················· | ···················· | ···················· | ···················· | ···················· | ···················· |
| X | 4741 | hCoV-19/England/ALDP-BC6584/2020\|EPI_ISL_721203\|20 | ···················· | ···················· | ···················· | ···················· | ···················· | ···················· |
| X | 4742 | hCoV-19/England/ALDP-BC77AB/2020\|EPI_ISL_721204\|20 | ···················· | ···················· | ···················· | ···················· | ···················· | ···················· |
| X | 4743 | hCoV-19/England/ALDP-BC76DB/2020\|EPI_ISL_721205\|20 | ···················· | ·········T·········· | ···················· | ···················· | ···················· | ···················· |
| X | 4744 | hCoV-19/England/ALDP-BC86BC/2020\|EPI_ISL_721217\|20 | ···················· | ···················· | ···················· | ···················· | ···················· | ···················· |
| X | 4745 | hCoV-19/England/ALDP-BC792D/2020\|EPI_ISL_721219\|20 | ···················· | ···················· | ···················· | ···················· | ···················· | ···················· |
| X | 4746 | hCoV-19/England/ALDP-BC6803/2020\|EPI_ISL_721220\|20 | ···················· | ···················· | ···················· | ···················· | ···················· | ···················· |
| X | 4747 | hCoV-19/England/ALDP-BC8519/2020\|EPI_ISL_721221\|20 | ···················· | ···················· | ···················· | ···················· | ···················· | ···················· |
| X | 4748 | hCoV-19/England/ALDP-BC7C7F/2020\|EPI_ISL_721222\|20 | ···················· | ···················· | ···················· | ···················· | ···················· | ···················· |
| X | 4749 | hCoV-19/England/ALDP-BC66AF/2020\|EPI_ISL_721223\|20 | ···················· | ···················· | ···················· | ···················· | ···················· | ···················· |
| X | 4750 | hCoV-19/England/ALDP-BC62D8/2020\|EPI_ISL_721224\|20 | ···················· | ···················· | ···················· | ···················· | ···················· | ···················· |
| X | 4751 | hCoV-19/England/ALDP-BC6575/2020\|EPI_ISL_721225\|20 | ···················· | ···················· | ···················· | ···················· | ···················· | ···················· |
| X | 4752 | hCoV-19/England/ALDP-BC7565/2020\|EPI_ISL_721226\|20 | ···················· | ···················· | ···················· | ···················· | ···················· | ···················· |
| X | 4753 | hCoV-19/England/ALDP-BC75ED/2020\|EPI_ISL_721227\|20 | ···················· | ···················· | ···················· | ···················· | ···················· | ···················· |
| X | 4754 | hCoV-19/England/ALDP-BC7AC0/2020\|EPI_ISL_721229\|20 | ···················· | ···················· | ···················· | ···················· | ···················· | ···················· |
| X | 4755 | hCoV-19/England/ALDP-BC62C9/2020\|EPI_ISL_721230\|20 | ···················· | ···················· | ···················· | ···················· | ···················· | ···················· |
| X | 4756 | hCoV-19/England/ALDP-BC8555/2020\|EPI_ISL_721231\|20 | ···················· | ···················· | ···················· | ···················· | ···················· | ···················· |
| X | 4757 | hCoV-19/England/ALDP-BC84C1/2020\|EPI_ISL_721233\|20 | ···················· | ···················· | ···················· | ···················· | ···················· | ···················· |
| X | 4758 | hCoV-19/England/ALDP-BC83B5/2020\|EPI_ISL_721234\|20 | ···················· | ···················· | ···················· | ···················· | ···················· | ···················· |
| X | 4759 | hCoV-19/England/ALDP-BC7D30/2020\|EPI_ISL_721235\|20 | ···················· | ···················· | ···················· | ···················· | ···················· | ···················· |
| X | 4760 | hCoV-19/England/ALDP-BC83A6/2020\|EPI_ISL_721236\|20 | ···················· | ···················· | ···················· | ···················· | ···················· | ···················· |
| X | 4761 | hCoV-19/England/ALDP-BC7AA2/2020\|EPI_ISL_721237\|20 | ···················· | ·A·················· | ···················· | ···················· | ···················· | ··A················· |
| X | 4762 | hCoV-19/England/ALDP-BC835B/2020\|EPI_ISL_721238\|20 | ···················· | ···················· | ···················· | ···················· | ···················· | ···················· |
| X | 4763 | hCoV-19/Netherlands/ZH-EMC-1072/2020\|EPI_ISL_72250 | ···················· | ···················· | ···················· | ···················· | ···················· | ···················· |
| X | 4764 | hCoV-19/Netherlands/ZH-EMC-921/2020\|EPI_ISL_722501 | ···················· | ···················· | ···················· | ···················· | ···················· | ···················· |
| X | 4765 | hCoV-19/Netherlands/DR-EMC-51/2020\|EPI_ISL_722502\| | ···················· | ···················· | ···················· | ···················· | ···················· | ···················· |
| X | 4766 | hCoV-19/Netherlands/ZH-EMC-1002/2020\|EPI_ISL_72250 | ···················· | ···················· | ···················· | ···················· | ···················· | ···················· |
| X | 4767 | hCoV-19/Netherlands/ZH-EMC-1073/2020\|EPI_ISL_72250 | ···················· | ···················· | ···················· | ···················· | ···················· | ···················· |
| X | 4768 | hCoV-19/Netherlands/ZH-EMC-1074/2020\|EPI_ISL_72250 | ···················· | ···················· | ···················· | ···················· | ···················· | ···················· |
| X | 4769 | hCoV-19/Netherlands/ZH-EMC-1075/2020\|EPI_ISL_72250 | ···················· | ···················· | ···················· | ···················· | ···················· | ···················· |
| X | 4770 | hCoV-19/Netherlands/ZH-EMC-931/2020\|EPI_ISL_722507 | ···················· | ···················· | ···················· | ···················· | ···················· | ···················· |
| X | 4771 | hCoV-19/Netherlands/NB-EMC-325/2020\|EPI_ISL_722508 | ···················· | ····A··············· | ···················· | ···················· | ···················· | ···················· |
| X | 4772 | hCoV-19/Netherlands/ZH-EMC-1076/2020\|EPI_ISL_72250 | ···················· | ···················· | ···················· | ···················· | ···················· | ···················· |
| X | 4773 | hCoV-19/Netherlands/ZH-EMC-961/2020\|EPI_ISL_722510 | ···················· | ···················· | ···················· | ···················· | ···················· | ···················· |
| X | 4774 | hCoV-19/Netherlands/GE-EMC-397/2020\|EPI_ISL_722511 | ···················· | ···················· | ···················· | ···················· | ···················· | ···················· |
| X | 4775 | hCoV-19/Netherlands/GE-EMC-399/2020\|EPI_ISL_722512 | ···················· | ···················· | ···················· | ···················· | ···················· | ···················· |
| X | 4776 | hCoV-19/Netherlands/ZH-EMC-945/2020\|EPI_ISL_722513 | ···················· | ···················· | ···················· | ···················· | ···················· | ···················· |
| X | 4777 | hCoV-19/Netherlands/UT-EMC-190/2020\|EPI_ISL_722514 | ···················· | ···················· | ···················· | ···················· | ···················· | ···················· |
| X | 4778 | hCoV-19/Netherlands/ZH-EMC-1077/2020\|EPI_ISL_72251 | ···················· | G··················· | ···················· | ···················· | ···················· | ···················· |
| X | 4779 | hCoV-19/Netherlands/ZH-EMC-979/2020\|EPI_ISL_722516 | ···················· | ···················· | ···················· | ···················· | ···················· | ···················· |
| X | 4780 | hCoV-19/Netherlands/ZH-EMC-929/2020\|EPI_ISL_722517 | ···················· | ···················· | ···················· | ···················· | ···················· | ···················· |
| X | 4781 | hCoV-19/Netherlands/UT-EMC-191/2020\|EPI_ISL_722518 | ···················· | ···················· | ···················· | ···················· | ···················· | ···················· |
| X | 4782 | hCoV-19/Netherlands/ZH-EMC-923/2020\|EPI_ISL_722519 | ···················· | ···················· | ···················· | ···················· | ···················· | ···················· |
| X | 4783 | hCoV-19/Netherlands/ZH-EMC-932/2020\|EPI_ISL_722520 | ···················· | ···················· | ···················· | ···················· | ···················· | ···················· |
| X | 4784 | hCoV-19/Netherlands/ZH-EMC-933/2020\|EPI_ISL_722521 | ···················· | ···················· | ···················· | ···················· | ···················· | ···················· |
| X | 4785 | hCoV-19/Netherlands/GE-EMC-356/2020\|EPI_ISL_722522 | ···················· | ···················· | ···················· | ···················· | ···················· | ···················· |
| X | 4786 | hCoV-19/Netherlands/GE-EMC-381/2020\|EPI_ISL_722523 | ···················· | ···················· | ···················· | ···················· | ···················· | ···················· |
| X | 4787 | hCoV-19/Netherlands/NB-EMC-344/2020\|EPI_ISL_722524 | ···················· | ···················· | ···················· | ···················· | ···················· | ···················· |
| X | 4788 | hCoV-19/Netherlands/NB-EMC-345/2020\|EPI_ISL_722525 | ···················· | ···················· | ···················· | ···················· | ···················· | ···················· |
| X | 4789 | hCoV-19/Netherlands/NB-EMC-329/2020\|EPI_ISL_722530 | ···················· | ···················· | ···················· | ···················· | ···················· | ···················· |
| X | 4790 | hCoV-19/Netherlands/NB-EMC-330/2020\|EPI_ISL_722531 | ···················· | ···················· | ···················· | ···················· | ···················· | ···················· |
| X | 4791 | hCoV-19/Netherlands/NB-EMC-351/2020\|EPI_ISL_722532 | ···················· | ···················· | ···················· | ···················· | ···················· | ···················· |
| X | 4792 | hCoV-19/Netherlands/NB-EMC-352/2020\|EPI_ISL_722533 | ···················· | ···················· | ···················· | ···················· | ···················· | ···················· |
| X | 4793 | hCoV-19/Netherlands/UT-EMC-192/2020\|EPI_ISL_722534 | ···················· | ···················· | ···················· | ···················· | ···················· | ···················· |
| X | 4794 | hCoV-19/Netherlands/UT-EMC-193/2020\|EPI_ISL_722535 | ···················· | ···················· | ···················· | ···················· | ···················· | ···················· |
| X | 4795 | hCoV-19/Netherlands/UT-EMC-208/2020\|EPI_ISL_722536 | ···················· | ···················· | ···················· | ···················· | ······T············· | ···················· |
| X | 4796 | hCoV-19/Netherlands/ZH-EMC-969/2020\|EPI_ISL_722548 | ···················· | ···················· | ···················· | ···················· | ···················· | ···················· |
| X | 4797 | hCoV-19/Netherlands/GE-EMC-420/2020\|EPI_ISL_722549 | ···················· | ···················· | ···················· | ···················· | ······T············· | ···················· |
| X | 4798 | hCoV-19/Netherlands/NB-EMC-336/2020\|EPI_ISL_722550 | ···················· | ···················· | ···················· | ···················· | ···················· | ···················· |
| X | 4799 | hCoV-19/Netherlands/NB-EMC-337/2020\|EPI_ISL_722551 | ···················· | ···················· | ···················· | ···················· | ···················· | ···················· |
| X | 4800 | hCoV-19/Netherlands/NB-EMC-338/2020\|EPI_ISL_722552 | ···················· | ···················· | ···················· | ···················· | ···················· | ···················· |
| X | 4801 | hCoV-19/Netherlands/NB-EMC-339/2020\|EPI_ISL_722553 | ···················· | ···················· | ···················· | ···················· | ···················· | ···················· |
| X | 4802 | hCoV-19/Netherlands/ZH-EMC-926/2020\|EPI_ISL_722554 | ···················· | ···················· | ···················· | ···················· | ···················· | ···················· |
| X | 4803 | hCoV-19/Netherlands/ZH-EMC-985/2020\|EPI_ISL_722555 | ···················· | ···················· | ···················· | ···················· | ···················· | ···················· |
| X | 4804 | hCoV-19/Netherlands/NB-EMC-353/2020\|EPI_ISL_722556 | ···················· | ···················· | ···················· | ···················· | ···················· | ···················· |
| X | 4805 | hCoV-19/Netherlands/NH-EMC-94/2020\|EPI_ISL_722557\| | ···················· | ···················· | ···················· | ···················· | ···················· | ···················· |
| X | 4806 | hCoV-19/Netherlands/UT-EMC-177/2020\|EPI_ISL_722558 | ···················· | ···················· | ···················· | ···················· | ···················· | ···················· |
| X | 4807 | hCoV-19/Netherlands/GE-EMC-357/2020\|EPI_ISL_722559 | ···················· | ···················· | ···················· | ···················· | ······T············· | ···················· |
| X | 4808 | hCoV-19/Netherlands/GE-EMC-395/2020\|EPI_ISL_722560 | ···················· | ···················· | ···················· | ···················· | ···················· | ···················· |
| X | 4809 | hCoV-19/Netherlands/NB-EMC-365/2020\|EPI_ISL_722561 | ···················· | ···················· | ···················· | ···················· | ···················· | ···················· |
| X | 4810 | hCoV-19/Netherlands/UT-EMC-183/2020\|EPI_ISL_722562 | ···················· | ···················· | ···················· | ···················· | ···················· | ···················· |
| X | 4811 | hCoV-19/Netherlands/GE-EMC-383/2020\|EPI_ISL_722563 | ···················· | ···················· | ···················· | ···················· | ···················· | ···················· |
| X | 4812 | hCoV-19/Netherlands/NB-EMC-340/2020\|EPI_ISL_722564 | ···················· | ···················· | ···················· | ···················· | ···················· | ···················· |
| X | 4813 | hCoV-19/Netherlands/NB-EMC-341/2020\|EPI_ISL_722565 | ···················· | ···················· | ···················· | ···················· | ···················· | ···················· |
| X | 4814 | hCoV-19/Netherlands/NB-EMC-342/2020\|EPI_ISL_722566 | ···················· | ···················· | ···················· | ···················· | ···················· | ···················· |
| X | 4815 | hCoV-19/Netherlands/NB-EMC-348/2020\|EPI_ISL_722567 | ···················· | ···················· | ···················· | ···················· | ···················· | ···················· |
| X | 4816 | hCoV-19/Netherlands/NB-EMC-354/2020\|EPI_ISL_722568 | ···················· | ···················· | ···················· | ···················· | ···················· | ···················· |
| X | 4817 | hCoV-19/Netherlands/ZH-EMC-953/2020\|EPI_ISL_722569 | ···················· | ···················· | ···················· | ···················· | ···················· | ···················· |
| X | 4818 | hCoV-19/England/ALDP-B76DD6/2020\|EPI_ISL_723813\|20 | ···················· | ·A·················· | ···················· | ···················· | ···················· | ··A················· |
| X | 4819 | hCoV-19/England/ALDP-B76DE5/2020\|EPI_ISL_723814\|20 | ···················· | ···················· | ···················· | ···················· | ···················· | ···················· |

| | | | | | | | |
|---|---|---|---|---|---|---|---|
| X | 4820 hCoV-19/England/ALDP-B76DF4/2020\|EPI_ISL_723815\|20 | ···················· | ···················· | ···················· | ···················· | ···················· | ···················· |
| X | 4821 hCoV-19/England/ALDP-B76E00/2020\|EPI_ISL_723816\|20 | ···················· | ···················· | ···················· | ···················· | ···················· | ···················· |
| X | 4822 hCoV-19/England/ALDP-B76E1F/2020\|EPI_ISL_723817\|20 | ···················· | ···················· | ···················· | ···················· | ···················· | ···················· |
| X | 4823 hCoV-19/England/ALDP-B76E2E/2020\|EPI_ISL_723818\|20 | ···················· | ···················· | ···················· | ···················· | ···················· | ···················· |
| X | 4824 hCoV-19/England/ALDP-B76E3D/2020\|EPI_ISL_723819\|20 | ···················· | ···················· | ···················· | ···················· | ···················· | ···················· |
| X | 4825 hCoV-19/England/ALDP-B76E4C/2020\|EPI_ISL_723820\|20 | ···················· | ···················· | ···················· | ···················· | ···················· | ···················· |
| X | 4826 hCoV-19/England/ALDP-B76E5B/2020\|EPI_ISL_723821\|20 | ···················· | ···················· | ···················· | ···················· | ···················· | ···················· |
| X | 4827 hCoV-19/England/ALDP-B76E79/2020\|EPI_ISL_723822\|20 | ···················· | ·········T·········· | ···················· | ···················· | ···················· | ···················· |
| X | 4828 hCoV-19/England/ALDP-B76EB5/2020\|EPI_ISL_723823\|20 | ···················· | ···················· | ···················· | ···················· | ···················· | ···················· |
| X | 4829 hCoV-19/England/ALDP-B76EC4/2020\|EPI_ISL_723824\|20 | ·T·················· | ···················· | ···················· | ···················· | ···················· | ···················· |
| X | 4830 hCoV-19/England/ALDP-B76ED3/2020\|EPI_ISL_723825\|20 | ···················· | ···················· | ···················· | ···················· | ···················· | ···················· |
| X | 4831 hCoV-19/England/ALDP-B76EF1/2020\|EPI_ISL_723826\|20 | ···················· | ···················· | ···················· | ···················· | ···················· | ···················· |
| X | 4832 hCoV-19/England/ALDP-B76F0D/2020\|EPI_ISL_723827\|20 | ···················· | ···················· | ···················· | ···················· | ···················· | ···················· |
| X | 4833 hCoV-19/Wales/ALDP-B76F1C/2020\|EPI_ISL_723828\|2020 | ···················· | ···················· | ···················· | ···················· | ···················· | ···················· |
| X | 4834 hCoV-19/England/ALDP-B76F2B/2020\|EPI_ISL_723829\|20 | ···················· | ···················· | ···················· | ···················· | ···················· | ···················· |
| X | 4835 hCoV-19/England/ALDP-B76F3A/2020\|EPI_ISL_723830\|20 | ···················· | ···················· | ···················· | ···················· | ···················· | ···················· |
| X | 4836 hCoV-19/England/ALDP-B76F58/2020\|EPI_ISL_723831\|20 | ···················· | ···················· | ···················· | ···················· | ···················· | ···················· |
| X | 4837 hCoV-19/England/ALDP-B76F76/2020\|EPI_ISL_723832\|20 | ···················· | ···················· | ···················· | ···················· | ···················· | ···················· |
| X | 4838 hCoV-19/England/ALDP-B76FA3/2020\|EPI_ISL_723833\|20 | ···················· | ···················· | ···················· | ···················· | ···················· | ···················· |
| X | 4839 hCoV-19/England/ALDP-B76FB2/2020\|EPI_ISL_723834\|20 | ···················· | ···················· | ···················· | ···················· | ···················· | ···················· |
| X | 4840 hCoV-19/England/ALDP-B76FC1/2020\|EPI_ISL_723835\|20 | ···················· | ···················· | ···················· | ···················· | ···················· | ···················· |
| X | 4841 hCoV-19/England/ALDP-B76FD0/2020\|EPI_ISL_723836\|20 | ···················· | ···················· | ···················· | ···················· | ···················· | ···················· |
| X | 4842 hCoV-19/England/ALDP-B76FFE/2020\|EPI_ISL_723837\|20 | ···················· | ···················· | ···················· | ···················· | ···················· | ···················· |
| X | 4843 hCoV-19/England/ALDP-B77009/2020\|EPI_ISL_723838\|20 | ···················· | ···················· | ···················· | ···················· | ···················· | ···················· |
| X | 4844 hCoV-19/England/ALDP-B77018/2020\|EPI_ISL_723839\|20 | ···················· | ···················· | ···················· | ···················· | ···················· | ···················· |
| X | 4845 hCoV-19/England/ALDP-B77054/2020\|EPI_ISL_723840\|20 | ···················· | ···················· | ···················· | ···················· | ···················· | ···················· |
| X | 4846 hCoV-19/England/ALDP-B77063/2020\|EPI_ISL_723841\|20 | ···················· | ···················· | ···················· | ···················· | ···················· | ···················· |
| X | 4847 hCoV-19/England/ALDP-B77081/2020\|EPI_ISL_723843\|20 | ···················· | ···················· | ···················· | ···················· | ···················· | ···················· |
| X | 4848 hCoV-19/England/ALDP-B77090/2020\|EPI_ISL_723844\|20 | ···················· | ···················· | ···················· | ···················· | ···················· | ···················· |
| X | 4849 hCoV-19/England/ALDP-B770FA/2020\|EPI_ISL_723845\|20 | ···················· | ···················· | ···················· | ···················· | ···················· | ···················· |
| X | 4850 hCoV-19/England/ALDP-B77160/2020\|EPI_ISL_723847\|20 | ···················· | ···················· | ···················· | ···················· | ···················· | ···················· |
| X | 4851 hCoV-19/England/ALDP-B7735B/2020\|EPI_ISL_723848\|20 | ···················· | ···················· | ···················· | ···················· | ···················· | ···················· |
| X | 4852 hCoV-19/England/ALDP-B7736A/2020\|EPI_ISL_723849\|20 | ···················· | ···················· | ···················· | ···················· | ···················· | ···················· |
| X | 4853 hCoV-19/England/ALDP-B773A6/2020\|EPI_ISL_723850\|20 | ···················· | ···················· | ···················· | ···················· | ···················· | ···················· |
| X | 4854 hCoV-19/England/CAMB-1BC196/2020\|EPI_ISL_723853\|20 | ···················· | ···················· | ···················· | ···················· | ···················· | ···················· |
| X | 4855 hCoV-19/England/CAMB-1BC1D2/2020\|EPI_ISL_723856\|20 | ···················· | ···················· | ···················· | ···················· | ···················· | ···················· |
| X | 4856 hCoV-19/England/CAMB-1BC2A2/2020\|EPI_ISL_723860\|20 | ···················· | ···················· | ···················· | ···················· | ······T············· | ···················· |
| X | 4857 hCoV-19/England/CAMB-1BC336/2020\|EPI_ISL_723863\|20 | ···················· | ···················· | ···················· | ···················· | ···················· | ···················· |
| X | 4858 hCoV-19/England/CAMB-1BC345/2020\|EPI_ISL_723864\|20 | ···················· | ···················· | ···················· | ···················· | ···················· | ···················· |
| X | 4859 hCoV-19/England/CAMB-1BC6A6/2020\|EPI_ISL_723886\|20 | ···················· | ···················· | ···················· | ···················· | ···················· | ···················· |
| X | 4860 hCoV-19/England/CAMB-1BC70D/2020\|EPI_ISL_723888\|20 | ···················· | ···················· | ···················· | ···················· | ···················· | ···················· |
| X | 4861 hCoV-19/England/CAMB-1BC785/2020\|EPI_ISL_723894\|20 | ···················· | ···················· | ···················· | ···················· | ···················· | ···················· |
| X | 4862 hCoV-19/England/CAMB-1BC794/2020\|EPI_ISL_723895\|20 | ···················· | ···················· | ···················· | ···················· | ···················· | ···················· |
| X | 4863 hCoV-19/England/CAMB-1BC7C1/2020\|EPI_ISL_723896\|20 | ···················· | ···················· | ···················· | ···················· | ······T············· | ···················· |
| X | 4864 hCoV-19/England/CAMB-1BCD92/2020\|EPI_ISL_723897\|20 | ···················· | ···················· | ···················· | ···················· | ······T············· | ···················· |
| X | 4865 hCoV-19/Croatia/Zagreb-19/2020\|EPI_ISL_710550\|2020 | ···················· | ···················· | ···················· | ···················· | ···················· | ···················· |
| X | 4866 hCoV-19/Croatia/Zagreb-20/2020\|EPI_ISL_710551\|2020 | ···················· | ···················· | ···················· | ···················· | ···················· | ···················· |
| X | 4867 hCoV-19/Croatia/Zagreb-24/2020\|EPI_ISL_710552\|2020 | ···················· | ···················· | ···················· | ···················· | ···················· | ···················· |
| X | 4868 hCoV-19/Croatia/Zagreb-9/2020\|EPI_ISL_710558\|2020- | ···················· | ···················· | ···················· | ···················· | ···················· | ···················· |
| X | 4869 hCoV-19/Croatia/Zagreb-16/2020\|EPI_ISL_710559\|2020 | ···················· | ···················· | ···················· | ···················· | ···················· | ···················· |
| X | 4870 hCoV-19/Croatia/Zagreb-6/2020\|EPI_ISL_710560\|2020- | ···················· | ···················· | ···················· | ···················· | ···················· | ···················· |
| X | 4871 hCoV-19/Croatia/Zagreb-17/2020\|EPI_ISL_710561\|2020 | ···················· | ···················· | ···················· | ···················· | ···················· | ···················· |
| X | 4872 hCoV-19/Croatia/Zagreb-18/2020\|EPI_ISL_710562\|2020 | ···················· | ···················· | ···················· | ···················· | ···················· | ···················· |
| X | 4873 hCoV-19/Croatia/Zagreb-28/2020\|EPI_ISL_710563\|2020 | ···················· | ···················· | ···················· | ···················· | ···················· | ···················· |
| X | 4874 hCoV-19/Croatia/Zagreb-23/2020\|EPI_ISL_710564\|2020 | ···················· | ···················· | ···················· | ···················· | ···················· | ···················· |
| X | 4875 hCoV-19/Croatia/Zagreb-25/2020\|EPI_ISL_710565\|2020 | ···················· | ···················· | ···················· | ···················· | ···················· | ···················· |
| X | 4876 hCoV-19/Croatia/Zagreb-21/2020\|EPI_ISL_710566\|2020 | ···················· | ···················· | ···················· | ···················· | ···················· | ···················· |
| X | 4877 hCoV-19/Croatia/Zagreb-2/2020\|EPI_ISL_710567\|2020- | ···················· | ···················· | ···················· | ···················· | ···················· | ···················· |
| X | 4878 hCoV-19/Croatia/Zagreb-10/2020\|EPI_ISL_710568\|2020 | ···················· | ···················· | ···················· | ···················· | ···················· | ···················· |
| X | 4879 hCoV-19/Sweden/20-53471/2020\|EPI_ISL_710590\|2020-1 | ···················· | ···················· | ···················· | ···················· | ···················· | ···················· |
| X | 4880 hCoV-19/Sweden/20-53631/2020\|EPI_ISL_710591\|2020-1 | ···················· | ···················· | ···················· | ···················· | ···················· | ···················· |
| X | 4881 hCoV-19/Sweden/20-53300/2020\|EPI_ISL_710592\|2020-1 | ···················· | ···················· | ···················· | ···················· | ···················· | ···················· |
| X | 4882 hCoV-19/Sweden/20-53306/2020\|EPI_ISL_710593\|2020-1 | ···················· | ···················· | ···················· | ···················· | ···················· | ···················· |
| X | 4883 hCoV-19/Sweden/20-52918/2020\|EPI_ISL_710594\|2020-0 | ···················· | ·A·················· | ···················· | ···················· | ···················· | ···A················ |
| X | 4884 hCoV-19/Sweden/20-08940/2020\|EPI_ISL_710595\|2020-0 | ···················· | ···················· | ···················· | ···················· | ···················· | ···················· |
| X | 4885 hCoV-19/Sweden/20-24259/2020\|EPI_ISL_710596\|2020-1 | ···················· | ······A············· | ···················· | ···················· | ···················· | ···················· |
| X | 4886 hCoV-19/Sweden/20-09859/2020\|EPI_ISL_710597\|2020-1 | ···················· | ···················· | ···················· | ···················· | ···················· | ···················· |
| X | 4887 hCoV-19/Sweden/20-53340/2020\|EPI_ISL_710598\|2020-1 | ···················· | ·A·················· | ···················· | ···················· | ···················· | ···················· |
| X | 4888 hCoV-19/Sweden/20-53341/2020\|EPI_ISL_710599\|2020-1 | ···················· | ···················· | ···················· | ···················· | ···················· | ···················· |
| X | 4889 hCoV-19/Sweden/20-53342/2020\|EPI_ISL_710600\|2020-1 | ···················· | ···················· | ···················· | ···················· | ···················· | ···················· |
| X | 4890 hCoV-19/Sweden/20-53343/2020\|EPI_ISL_710601\|2020-1 | ···················· | ·A·················· | ···················· | ···················· | ···················· | ···················· |
| X | 4891 hCoV-19/Sweden/20-53344/2020\|EPI_ISL_710602\|2020-1 | ···················· | ·A·················· | ···················· | ···················· | ···················· | ···A················ |
| X | 4892 hCoV-19/Sweden/20-53635/2020\|EPI_ISL_710603\|2020-1 | ···················· | ···················· | ···················· | ···················· | ···················· | ···················· |
| X | 4893 hCoV-19/Sweden/20-53637/2020\|EPI_ISL_710604\|2020-1 | ···················· | ·A·················· | ···················· | ···················· | ···················· | ···················· |
| X | 4894 hCoV-19/Sweden/20-53022/2020\|EPI_ISL_710605\|2020-0 | ···················· | ·A·················· | ···················· | ···················· | ···················· | ···················· |
| X | 4895 hCoV-19/Sweden/20-53366/2020\|EPI_ISL_710606\|2020-1 | ···················· | ···················· | ···················· | ···················· | ···················· | ···················· |
| X | 4896 hCoV-19/Sweden/20-53445/2020\|EPI_ISL_710607\|2020-1 | ···················· | ···················· | ···················· | ···················· | ···················· | ···················· |
| X | 4897 hCoV-19/Sweden/20-52811/2020\|EPI_ISL_710608\|2020-0 | ···················· | ···················· | ···················· | ···················· | ···················· | ···················· |
| X | 4898 hCoV-19/Sweden/20-53057/2020\|EPI_ISL_710609\|2020-0 | ···················· | ···G················ | ···················· | ···················· | ···················· | ···················· |
| X | 4899 hCoV-19/Sweden/20-53179/2020\|EPI_ISL_710610\|2020-1 | ···················· | ···················· | ···················· | ···················· | ······T············· | ···················· |

X 4900 hCoV-19/Sweden/20-53248/2020|EPI_ISL_710611|2020-1 ···················· ·A·················· ···················· ···················· ···················· ····················
X 4901 hCoV-19/Sweden/20-24688/2020|EPI_ISL_710612|2020-1 ···················· ···············G···· ···················· ···················· ···················· ····················
X 4902 hCoV-19/Sweden/20-53337/2020|EPI_ISL_710613|2020-1 ···················· ···················· ···················· ···················· ···················· ····················
X 4903 hCoV-19/Wales/ALDP-BE8E81/2020|EPI_ISL_710614|2020 ···················· ···················· ···················· ···················· ···················· ····················
X 4904 hCoV-19/Scotland/QEUH-BEAE61/2020|EPI_ISL_710615|2 ···················· ·A·················· ···················· ···················· ···················· ···A················
X 4905 hCoV-19/Wales/ALDP-BE95F4/2020|EPI_ISL_710616|2020 ···················· ···················· ···················· ···················· ···················· ····················
X 4906 hCoV-19/Wales/ALDP-BE95E5/2020|EPI_ISL_710617|2020 ···················· ···················· ···················· ···················· ···················· ····················
X 4907 hCoV-19/Scotland/QEUH-BEB1D0/2020|EPI_ISL_710619|2 ···················· ···················· ···················· ···················· ···················· ····················
X 4908 hCoV-19/Wales/ALDP-BE9257/2020|EPI_ISL_710620|2020 ···················· ···················· ···················· ···················· ···················· ····················
X 4909 hCoV-19/Scotland/QEUH-BEAE9E/2020|EPI_ISL_710621|2 ···················· ···················· ···················· ···················· ···················· ····················
X 4910 hCoV-19/Scotland/QEUH-BEAEF8/2020|EPI_ISL_710622|2 ···················· ···················· ···················· ···················· ···················· ····················
X 4911 hCoV-19/Scotland/QEUH-BEAE70/2020|EPI_ISL_710623|2 ···················· ···················· ···················· ···················· ···················· ····················
X 4912 hCoV-19/Denmark/DCGC-14631/2020|EPI_ISL_711880|202 ···················· ···················· ···················· ···················· ···················· ····················
X 4913 hCoV-19/Denmark/DCGC-14632/2020|EPI_ISL_711881|202 ···················· ···················· ···················· ···················· ···················· ····················
X 4914 hCoV-19/Denmark/DCGC-14633/2020|EPI_ISL_711882|202 ···················· ···················· ···················· ···················· ···················· ····················
X 4915 hCoV-19/Denmark/DCGC-14636/2020|EPI_ISL_711884|202 ···················· ···················· ···················· ···················· ···················· ····················
X 4916 hCoV-19/Denmark/DCGC-14648/2020|EPI_ISL_711889|202 ···················· ·A·················· ···················· ···················· ···················· ····················
X 4917 hCoV-19/Denmark/DCGC-14650/2020|EPI_ISL_711890|202 ···················· ···················· ···················· ···················· ···················· ····················
X 4918 hCoV-19/Denmark/DCGC-14651/2020|EPI_ISL_711891|202 ···················· ···················· ···················· ···················· ···················· ····················
X 4919 hCoV-19/Denmark/DCGC-14652/2020|EPI_ISL_711892|202 ···················· ···················· ···················· ···················· ···················· ····················
X 4920 hCoV-19/Denmark/DCGC-14653/2020|EPI_ISL_711893|202 ···················· ···················· ···················· ···················· ···················· ····················
X 4921 hCoV-19/Denmark/DCGC-14654/2020|EPI_ISL_711894|202 ···················· ···················· ···················· ···················· ···················· ····················
X 4922 hCoV-19/Denmark/DCGC-14658/2020|EPI_ISL_711895|202 ···················· ···················· ···················· ···················· ···················· ····················
X 4923 hCoV-19/Denmark/DCGC-14660/2020|EPI_ISL_711896|202 ···················· ···················· ···················· ···················· ···················· ····················
X 4924 hCoV-19/Denmark/DCGC-14664/2020|EPI_ISL_711897|202 ···················· ···················· ···················· ···················· ···················· ····················
X 4925 hCoV-19/Denmark/DCGC-14666/2020|EPI_ISL_711898|202 ···················· ···················· ···················· ···················· ···················· ····················
X 4926 hCoV-19/Denmark/DCGC-14667/2020|EPI_ISL_711899|202 ···················· ···················· ···················· ···················· ···················· ····················
X 4927 hCoV-19/Denmark/DCGC-14668/2020|EPI_ISL_711900|202 ···················· ···················· ···················· ···················· ···················· ····················
X 4928 hCoV-19/Denmark/DCGC-14669/2020|EPI_ISL_711901|202 ···················· ···················· ···················· ···················· ···················· ····················
X 4929 hCoV-19/Denmark/DCGC-14670/2020|EPI_ISL_711902|202 ···················· ···················· ···················· ···················· ···················· ····················
X 4930 hCoV-19/Denmark/DCGC-14674/2020|EPI_ISL_711903|202 ···················· ···················· ···················· ···················· ···················· ····················
X 4931 hCoV-19/Denmark/DCGC-14675/2020|EPI_ISL_711904|202 ···················· ···················· ···················· ···················· ···················· ····················
X 4932 hCoV-19/Denmark/DCGC-14676/2020|EPI_ISL_711905|202 ···················· ···················· ···················· ···················· ···················· ····················
X 4933 hCoV-19/Denmark/DCGC-14679/2020|EPI_ISL_711906|202 ···················· ···················· ···················· ···················· ···················· ····················
X 4934 hCoV-19/Denmark/DCGC-14681/2020|EPI_ISL_711907|202 ···················· ···················· ···················· ···················· ···················· ····················
X 4935 hCoV-19/Denmark/DCGC-14682/2020|EPI_ISL_711908|202 ···················· ···················· ···················· ···················· ···················· ····················
X 4936 hCoV-19/Denmark/DCGC-14684/2020|EPI_ISL_711909|202 ···················· ···················· ···················· ···················· ···················· ····················
X 4937 hCoV-19/Denmark/DCGC-14685/2020|EPI_ISL_711910|202 ···················· ···················· ···················· ···················· ···················· ····················
X 4938 hCoV-19/Denmark/DCGC-14686/2020|EPI_ISL_711911|202 ···················· ···················· ···················· ···················· ···················· ····················
X 4939 hCoV-19/Denmark/DCGC-14691/2020|EPI_ISL_711912|202 ···················· ···················· ···················· ···················· ···················· ····················
X 4940 hCoV-19/Denmark/DCGC-14692/2020|EPI_ISL_711913|202 ···················· ···················· ···················· ···················· ···················· ····················
X 4941 hCoV-19/Denmark/DCGC-14693/2020|EPI_ISL_711914|202 ···················· ···················· ···················· ···················· ···················· ····················
X 4942 hCoV-19/Denmark/DCGC-14698/2020|EPI_ISL_711915|202 ···················· ·A·················· ···················· ···················· ···················· ····················
X 4943 hCoV-19/Denmark/DCGC-14699/2020|EPI_ISL_711916|202 ···················· ···················· ···················· ···················· ···················· ····················
X 4944 hCoV-19/Denmark/DCGC-14700/2020|EPI_ISL_711917|202 ···················· ···················· ···················· ···················· ···················· ····················
X 4945 hCoV-19/Denmark/DCGC-14704/2020|EPI_ISL_711918|202 ···················· ···················· ···················· ···················· ···················· ····················
X 4946 hCoV-19/Denmark/DCGC-14705/2020|EPI_ISL_711919|202 ···················· ···················· ···················· ···················· ···················· ····················
X 4947 hCoV-19/Denmark/DCGC-14706/2020|EPI_ISL_711920|202 ···················· ···················· ···················· ···················· ···················· ····················
X 4948 hCoV-19/Denmark/DCGC-14707/2020|EPI_ISL_711921|202 ···················· ···················· ···················· ···················· ···················· ····················
X 4949 hCoV-19/Denmark/DCGC-14709/2020|EPI_ISL_711922|202 ···················· ···················· ···················· ···················· ···················· ····················
X 4950 hCoV-19/Denmark/DCGC-14710/2020|EPI_ISL_711923|202 ···················· ···················· ···················· ···················· ···················· ····················
X 4951 hCoV-19/Denmark/DCGC-14712/2020|EPI_ISL_711924|202 ···················· ···················· ···················· ···················· ···················· ····················
X 4952 hCoV-19/Denmark/DCGC-14714/2020|EPI_ISL_711925|202 ···················· ···················· ···················· ···················· ···················· ····················
X 4953 hCoV-19/Denmark/DCGC-14715/2020|EPI_ISL_711926|202 ···················· ···················· ···················· ···················· ···················· ····················
X 4954 hCoV-19/Denmark/DCGC-14716/2020|EPI_ISL_711927|202 ···················· ···················· ···················· ···················· ···················· ····················
X 4955 hCoV-19/Denmark/DCGC-14717/2020|EPI_ISL_711928|202 ···················· ···················· ···················· ···················· ···················· ····················
X 4956 hCoV-19/Denmark/DCGC-14718/2020|EPI_ISL_711929|202 ···················· ···················· ···················· ···················· ···················· ····················
X 4957 hCoV-19/Denmark/DCGC-14720/2020|EPI_ISL_711930|202 ···················· ···················· ···················· ···················· ···················· ····················
X 4958 hCoV-19/Denmark/DCGC-14721/2020|EPI_ISL_711931|202 ···················· ···················· ···················· ···················· ···················· ····················
X 4959 hCoV-19/Denmark/DCGC-14723/2020|EPI_ISL_711932|202 ···················· ···················· ···················· ···················· ···················· ····················
X 4960 hCoV-19/Denmark/DCGC-14724/2020|EPI_ISL_711933|202 ···················· ···················· ···················· ···················· ···················· ····················
X 4961 hCoV-19/Denmark/DCGC-14725/2020|EPI_ISL_711934|202 ···················· ···················· ···················· ···················· ···················· ····················
X 4962 hCoV-19/Denmark/DCGC-14726/2020|EPI_ISL_711935|202 ···················· ···················· ···················· ···················· ···················· ····················
X 4963 hCoV-19/Denmark/DCGC-14728/2020|EPI_ISL_711936|202 ···················· ·A·················· ···················· ···················· ···················· ···A················
X 4964 hCoV-19/Denmark/DCGC-14729/2020|EPI_ISL_711937|202 ···················· ···················· ···················· ···················· ···················· ····················
X 4965 hCoV-19/Denmark/DCGC-14730/2020|EPI_ISL_711938|202 ···················· ···················· ···················· ···················· ···················· ····················
X 4966 hCoV-19/Denmark/DCGC-14731/2020|EPI_ISL_711939|202 ···················· ···················· ···················· ···················· ···················· ····················
X 4967 hCoV-19/Denmark/DCGC-14732/2020|EPI_ISL_711940|202 ···················· ···················· ···················· ···················· ···················· ····················
X 4968 hCoV-19/Denmark/DCGC-14733/2020|EPI_ISL_711941|202 ···················· ···················· ···················· ···················· ···················· ····················
X 4969 hCoV-19/Denmark/DCGC-14734/2020|EPI_ISL_711942|202 ···················· ···················· ···················· ···················· ···················· ····················
X 4970 hCoV-19/Denmark/DCGC-14735/2020|EPI_ISL_711943|202 ···················· ···················· ···················· ···················· ···················· ····················
X 4971 hCoV-19/Denmark/DCGC-14736/2020|EPI_ISL_711944|202 ···················· ···················· ···················· ···················· ···················· ····················
X 4972 hCoV-19/Denmark/DCGC-14738/2020|EPI_ISL_711945|202 ···················· ···················· ···················· ···················· ···················· ····················
X 4973 hCoV-19/Denmark/DCGC-14739/2020|EPI_ISL_711946|202 ···················· ···················· ···················· ···················· ···················· ····················
X 4974 hCoV-19/Denmark/DCGC-14740/2020|EPI_ISL_711947|202 ···················· ···················· ···················· ···················· ···················· ····················
X 4975 hCoV-19/Denmark/DCGC-14742/2020|EPI_ISL_711948|202 ···················· ·A·················· ···················· ···················· ···················· ····················
X 4976 hCoV-19/Denmark/DCGC-14743/2020|EPI_ISL_711949|202 ···················· ···················· ···················· ···················· ···················· ····················
X 4977 hCoV-19/Denmark/DCGC-14744/2020|EPI_ISL_711950|202 ···················· ···················· ···················· ···················· ···················· ····················
X 4978 hCoV-19/Denmark/DCGC-14745/2020|EPI_ISL_711951|202 ···················· ···················· ···················· ···················· ···················· ····················
X 4979 hCoV-19/Denmark/DCGC-14746/2020|EPI_ISL_711952|202 ···················· ···················· ···················· ···················· ···················· ····················

| | | | | | | | | |
|---|---|---|---|---|---|---|---|---|
| X | 4980 | hCoV-19/Denmark/DCGC-14747/2020\|EPI_ISL_711953\|202 | ···················· | ···················· | ···················· | ···················· | ···················· | ···················· |
| X | 4981 | hCoV-19/Denmark/DCGC-14749/2020\|EPI_ISL_711954\|202 | ···················· | ···················· | ···················· | ···················· | ···················· | ···················· |
| X | 4982 | hCoV-19/England/ALDP-BC8704/2020\|EPI_ISL_721206\|20 | ···················· | ···················· | ···················· | ···················· | ···················· | ···················· |
| X | 4983 | hCoV-19/England/ALDP-BC8537/2020\|EPI_ISL_721207\|20 | ···················· | ···················· | ···················· | ···················· | ···················· | ···················· |
| X | 4984 | hCoV-19/England/ALDP-BC84D0/2020\|EPI_ISL_721208\|20 | ···················· | ···················· | ···················· | ···················· | ···················· | ···················· |
| X | 4985 | hCoV-19/England/ALDP-BC788A/2020\|EPI_ISL_721209\|20 | ···················· | ···················· | ···················· | ···················· | ···················· | ···················· |
| X | 4986 | hCoV-19/England/ALDP-BC7705/2020\|EPI_ISL_721210\|20 | ···················· | ·A·················· | ···················· | ···················· | ···················· | ···A················ |
| X | 4987 | hCoV-19/England/ALDP-BC73E3/2020\|EPI_ISL_721211\|20 | ···················· | ···················· | ···················· | ···················· | ···················· | ···················· |
| X | 4988 | hCoV-19/England/ALDP-BC7617/2020\|EPI_ISL_721212\|20 | ···················· | ···················· | ···················· | ···················· | ···················· | ···················· |
| X | 4989 | hCoV-19/England/ALDP-BC83D3/2020\|EPI_ISL_721214\|20 | ···················· | ···················· | ···················· | ···················· | ···················· | ···················· |
| X | 4990 | hCoV-19/England/ALDP-BC8652/2020\|EPI_ISL_721215\|20 | ···················· | ···················· | ···················· | ···················· | ···················· | ···················· |
| X | 4991 | hCoV-19/England/ALDP-BC79F0/2020\|EPI_ISL_721216\|20 | ···················· | ·A·················· | ···················· | ···················· | ···················· | ···A········G······· |
| X | 4992 | hCoV-19/England/ALDP-BC7A39/2020\|EPI_ISL_721239\|20 | ···················· | ···················· | ···················· | ···················· | ···················· | ···················· |
| X | 4993 | hCoV-19/England/ALDP-BC79E1/2020\|EPI_ISL_721241\|20 | ···················· | ···················· | ···················· | ···················· | ···················· | ···················· |
| X | 4994 | hCoV-19/England/ALDP-BC7A66/2020\|EPI_ISL_721242\|20 | ···················· | ···················· | ···················· | ···················· | ···················· | ···················· |
| X | 4995 | hCoV-19/England/ALDP-BC638A/2020\|EPI_ISL_721243\|20 | ···················· | ···················· | ···················· | ···················· | ···················· | ···················· |
| X | 4996 | hCoV-19/England/ALDP-BC7DB8/2020\|EPI_ISL_721244\|20 | ···················· | ···················· | ···················· | ···················· | ···················· | ···················· |
| X | 4997 | hCoV-19/England/ALDP-BC778D/2020\|EPI_ISL_721245\|20 | ···················· | ···················· | ···················· | ···················· | ···················· | ···················· |
| X | 4998 | hCoV-19/England/ALDP-BC8A1A/2020\|EPI_ISL_721246\|20 | ···················· | ···················· | ···················· | ···················· | ······T············· | ···················· |
| X | 4999 | hCoV-19/England/ALDP-BC64A5/2020\|EPI_ISL_721247\|20 | ···················· | ···················· | ···················· | ···················· | ···················· | ···················· |
| X | 5000 | hCoV-19/England/ALDP-BC6821/2020\|EPI_ISL_721248\|20 | ···················· | ···················· | ···················· | ···················· | ···················· | ···················· |
| X | 5001 | hCoV-19/England/ALDP-BC684F/2020\|EPI_ISL_721249\|20 | ···················· | ···················· | ···················· | ···················· | ···················· | ···················· |
| X | 5002 | hCoV-19/England/ALDP-BC75FC/2020\|EPI_ISL_721251\|20 | ···················· | ···················· | ···················· | ···················· | ···················· | ···················· |
| X | 5003 | hCoV-19/England/ALDP-BC783F/2020\|EPI_ISL_721252\|20 | ···················· | ···················· | ···················· | ···················· | ···················· | ···················· |
| X | 5004 | hCoV-19/England/ALDP-BC89A4/2020\|EPI_ISL_721253\|20 | ···················· | ···················· | ···················· | ···················· | ···················· | ···················· |
| X | 5005 | hCoV-19/England/ALDP-BC637B/2020\|EPI_ISL_721255\|20 | ···················· | ···················· | ···················· | ···················· | ···················· | ···················· |
| X | 5006 | hCoV-19/England/ALDP-BC6645/2020\|EPI_ISL_721256\|20 | ···················· | ·········T·········· | ···················· | ···················· | ···················· | ···················· |
| X | 5007 | hCoV-19/England/ALDP-BC8810/2020\|EPI_ISL_721257\|20 | ···················· | ···················· | ···················· | ···················· | ···················· | ···················· |
| X | 5008 | hCoV-19/England/ALDP-BC6205/2020\|EPI_ISL_721259\|20 | ···················· | ···················· | ···················· | ···················· | ···················· | ···················· |
| X | 5009 | hCoV-19/England/ALDP-BC751A/2020\|EPI_ISL_721260\|20 | ···················· | ···················· | ···················· | ···················· | ···················· | ···················· |
| X | 5010 | hCoV-19/England/ALDP-BC77C9/2020\|EPI_ISL_721261\|20 | ···················· | ···················· | ···················· | ···················· | ···················· | ···················· |
| X | 5011 | hCoV-19/England/ALDP-BC7459/2020\|EPI_ISL_721262\|20 | ···················· | ···················· | ···················· | ···················· | ···················· | ···················· |
| X | 5012 | hCoV-19/England/ALDP-BC75B0/2020\|EPI_ISL_721263\|20 | ···················· | ···················· | ···················· | ···················· | ···················· | ···················· |
| X | 5013 | hCoV-19/England/ALDP-BC7662/2020\|EPI_ISL_721264\|20 | ···················· | ···················· | ···················· | ···················· | ······T············· | ···················· |
| X | 5014 | hCoV-19/England/ALDP-BC877D/2020\|EPI_ISL_721266\|20 | ···················· | ···················· | ···················· | ···················· | ···················· | ···················· |
| X | 5015 | hCoV-19/England/ALDP-BC7BAF/2020\|EPI_ISL_721267\|20 | ···················· | ···················· | ···················· | ···················· | ···················· | ···················· |
| X | 5016 | hCoV-19/England/ALDP-BC7732/2020\|EPI_ISL_721268\|20 | ···················· | ···················· | ···················· | ···················· | ···················· | ···················· |
| X | 5017 | hCoV-19/England/ALDP-BC8546/2020\|EPI_ISL_721269\|20 | ···················· | ···················· | ···················· | ···················· | ···················· | ···················· |
| X | 5018 | hCoV-19/England/ALDP-BC6618/2020\|EPI_ISL_721270\|20 | ···················· | ·A·················· | ···················· | ···················· | ···················· | ···A················ |
| X | 5019 | hCoV-19/England/ALDP-BC645A/2020\|EPI_ISL_721271\|20 | ···················· | ···················· | ···················· | ···················· | ···················· | ···················· |
| X | 5020 | hCoV-19/England/ALDP-BC6715/2020\|EPI_ISL_721273\|20 | ···················· | ···················· | ···················· | ···················· | ···················· | ···················· |
| X | 5021 | hCoV-19/England/ALDP-BC7D03/2020\|EPI_ISL_721274\|20 | ···················· | ···················· | ···················· | ···················· | ···················· | ···················· |
| X | 5022 | hCoV-19/England/ALDP-BC61AE/2020\|EPI_ISL_721275\|20 | ···················· | ···················· | ···················· | ···················· | ···················· | ···················· |
| X | 5023 | hCoV-19/England/ALDP-BC65C0/2020\|EPI_ISL_721276\|20 | ···················· | ···················· | ···················· | ···················· | ···················· | ···················· |
| X | 5024 | hCoV-19/England/ALDP-BC7D8B/2020\|EPI_ISL_721277\|20 | ···················· | ···················· | ···················· | ···················· | ···················· | ···················· |
| X | 5025 | hCoV-19/England/ALDP-BC8582/2020\|EPI_ISL_721278\|20 | ···················· | ···················· | ···················· | ···················· | ···················· | ···················· |
| X | 5026 | hCoV-19/England/ALDP-BC65EE/2020\|EPI_ISL_721279\|20 | ···················· | ···················· | ···················· | ···················· | ···················· | ···················· |
| X | 5027 | hCoV-19/England/ALDP-BC6627/2020\|EPI_ISL_721280\|20 | ···················· | ···················· | ···················· | ···················· | ···················· | ···················· |
| X | 5028 | hCoV-19/England/ALDP-BC642D/2020\|EPI_ISL_721281\|20 | ···················· | ···················· | ···················· | ···················· | ···················· | ···················· |
| X | 5029 | hCoV-19/England/ALDP-BC85A0/2020\|EPI_ISL_721282\|20 | ···················· | ···················· | ···················· | ···················· | ···················· | ···················· |
| X | 5030 | hCoV-19/England/ALDP-BC7B18/2020\|EPI_ISL_721283\|20 | ···················· | ···················· | ···················· | ···················· | ···················· | ···················· |
| X | 5031 | hCoV-19/England/ALDP-BC634E/2020\|EPI_ISL_721284\|20 | ···················· | ···················· | ···················· | ···················· | ···················· | ···················· |
| X | 5032 | hCoV-19/England/ALDP-BC89FF/2020\|EPI_ISL_721286\|20 | ···················· | ···················· | ···················· | ···················· | ···················· | ···················· |
| X | 5033 | hCoV-19/England/ALDP-BC62BA/2020\|EPI_ISL_721287\|20 | ···················· | ···················· | ···················· | ···················· | ···················· | ···················· |
| X | 5034 | hCoV-19/England/ALDP-BC883E/2020\|EPI_ISL_721288\|20 | ···················· | ···················· | ···················· | ···················· | ···················· | ···A················ |
| X | 5035 | hCoV-19/England/ALDP-BC7BCD/2020\|EPI_ISL_721289\|20 | ···················· | ···················· | ···················· | ···················· | ···················· | ···················· |
| X | 5036 | hCoV-19/England/ALDP-BC79C3/2020\|EPI_ISL_721290\|20 | ···················· | ···················· | ···················· | ···················· | ···················· | ···················· |
| X | 5037 | hCoV-19/England/ALDP-BC742C/2020\|EPI_ISL_721291\|20 | ···················· | ···················· | ···················· | ···················· | ···················· | ···················· |
| X | 5038 | hCoV-19/England/ALDP-BC87C8/2020\|EPI_ISL_721292\|20 | ···················· | ···················· | ···················· | ···················· | ···················· | ···················· |
| X | 5039 | hCoV-19/England/ALDP-BC7592/2020\|EPI_ISL_721293\|20 | ···················· | ···················· | ···················· | ···················· | ···················· | ···················· |
| X | 5040 | hCoV-19/England/ALDP-BC7C42/2020\|EPI_ISL_721294\|20 | ···················· | ···················· | ···················· | ···················· | ···················· | ···················· |
| X | 5041 | hCoV-19/England/ALDP-BC7C33/2020\|EPI_ISL_721295\|20 | ···················· | ···················· | ···················· | ···················· | ···················· | ···················· |
| X | 5042 | hCoV-19/England/ALDP-BC785D/2020\|EPI_ISL_721296\|20 | ···················· | ···················· | ···················· | ···················· | ···················· | ···················· |
| X | 5043 | hCoV-19/England/ALDP-BC76F9/2020\|EPI_ISL_721297\|20 | ···················· | ···················· | ···················· | ···················· | ···················· | ···················· |
| X | 5044 | hCoV-19/England/ALDP-BC6478/2020\|EPI_ISL_721298\|20 | ···················· | ···················· | ···················· | ···················· | ···················· | ···················· |
| X | 5045 | hCoV-19/England/ALDP-BC7750/2020\|EPI_ISL_721299\|20 | ···················· | ···················· | ···················· | ···················· | ···················· | ···················· |
| X | 5046 | hCoV-19/Netherlands/NB-EMC-346/2020\|EPI_ISL_722526 | ···················· | ···················· | ···················· | ···················· | ···················· | ···················· |
| X | 5047 | hCoV-19/Netherlands/OV-EMC-88/2020\|EPI_ISL_722527\| | ···················· | ···················· | ···················· | ···················· | ······T············· | ···················· |
| X | 5048 | hCoV-19/Netherlands/NB-EMC-326/2020\|EPI_ISL_722528 | ···················· | ···················· | ···················· | ···················· | ···················· | ···················· |
| X | 5049 | hCoV-19/Netherlands/NB-EMC-471/2020\|EPI_ISL_722529 | ···················· | ···················· | ···················· | ···················· | ···················· | ···················· |
| X | 5050 | hCoV-19/Netherlands/GE-EMC-382/2020\|EPI_ISL_722537 | ···················· | ···················· | ···················· | ···················· | ···················· | ···················· |
| X | 5051 | hCoV-19/Netherlands/NB-EMC-332/2020\|EPI_ISL_722538 | ···················· | ····A··············· | ···················· | ···················· | ···················· | ···················· |
| X | 5052 | hCoV-19/Netherlands/NB-EMC-334/2020\|EPI_ISL_722539 | ···················· | ····A··············· | ···················· | ···················· | ···················· | ···················· |
| X | 5053 | hCoV-19/Netherlands/NB-EMC-335/2020\|EPI_ISL_722540 | ···················· | ···················· | ···················· | ···················· | ···················· | ···················· |
| X | 5054 | hCoV-19/Netherlands/NB-EMC-357/2020\|EPI_ISL_722541 | ···················· | ···················· | ···················· | ···················· | ···················· | ···················· |
| X | 5055 | hCoV-19/Netherlands/NB-EMC-358/2020\|EPI_ISL_722542 | ···················· | ···················· | ···················· | ···················· | ···················· | ···················· |
| X | 5056 | hCoV-19/Netherlands/NH-EMC-93/2020\|EPI_ISL_722543\| | ···················· | ···················· | ···················· | ···················· | ···················· | ···················· |
| X | 5057 | hCoV-19/Netherlands/ZH-EMC-934/2020\|EPI_ISL_722544 | ···················· | ···················· | ···················· | ···················· | ······T············· | ···················· |
| X | 5058 | hCoV-19/Netherlands/GE-EMC-400/2020\|EPI_ISL_722545 | ···················· | ···················· | ···················· | ···················· | ···················· | ···················· |
| X | 5059 | hCoV-19/Netherlands/GE-EMC-403/2020\|EPI_ISL_722546 | ···················· | ···················· | ···················· | ···················· | ···················· | ···················· |

| | | | | | | | |
|---|---|---|---|---|---|---|---|
| X | 5060 hCoV-19/Netherlands/ZH-EMC-1049/2020\|EPI_ISL_72254 | ···················· | ···················· | ···················· | ···················· | ···················· | ···················· |
| X | 5061 hCoV-19/Netherlands/GE-EMC-369/2020\|EPI_ISL_722570 | ···················· | ···················· | ···················· | ···················· | ···················· | ···················· |
| X | 5062 hCoV-19/Netherlands/GE-EMC-404/2020\|EPI_ISL_722571 | ···················· | ···················· | ···················· | ···················· | ···················· | ···················· |
| X | 5063 hCoV-19/Netherlands/LI-EMC-102/2020\|EPI_ISL_722572 | ············T······· | ···················· | ···················· | ···················· | ···················· | ···················· |
| X | 5064 hCoV-19/Netherlands/LI-EMC-103/2020\|EPI_ISL_722573 | ············T······· | ···················· | ···················· | ···················· | ···················· | ···················· |
| X | 5065 hCoV-19/Netherlands/LI-EMC-104/2020\|EPI_ISL_722574 | ············T······· | ···················· | ···················· | ···················· | ···················· | ···················· |
| X | 5066 hCoV-19/Netherlands/LI-EMC-105/2020\|EPI_ISL_722575 | ············T······· | ···················· | ···················· | ···················· | ···················· | ···················· |
| X | 5067 hCoV-19/Netherlands/LI-EMC-106/2020\|EPI_ISL_722576 | ············T······· | ···················· | ···················· | ···················· | ···················· | ···················· |
| X | 5068 hCoV-19/Netherlands/LI-EMC-107/2020\|EPI_ISL_722577 | ············T······· | ···················· | ···················· | ···················· | ···················· | ···················· |
| X | 5069 hCoV-19/Netherlands/LI-EMC-110/2020\|EPI_ISL_722578 | ············T······· | ···················· | ···················· | ···················· | ···················· | ···················· |
| X | 5070 hCoV-19/Netherlands/LI-EMC-111/2020\|EPI_ISL_722579 | ············T······· | ···················· | ···················· | ···················· | ···················· | ···················· |
| X | 5071 hCoV-19/Netherlands/UT-EMC-157/2020\|EPI_ISL_722580 | ···················· | ···················· | ···················· | ···················· | ···················· | ···················· |
| X | 5072 hCoV-19/Netherlands/GE-EMC-394/2020\|EPI_ISL_722581 | ···················· | ···················· | ···················· | ···················· | ···················· | ···················· |
| X | 5073 hCoV-19/Netherlands/LI-EMC-108/2020\|EPI_ISL_722582 | ············T······· | ···················· | ···················· | ···················· | ···················· | ···················· |
| X | 5074 hCoV-19/Netherlands/NB-EMC-343/2020\|EPI_ISL_722583 | ···················· | ···················· | ···················· | ···················· | ···················· | ···················· |
| X | 5075 hCoV-19/Netherlands/NB-EMC-349/2020\|EPI_ISL_722584 | ···················· | ···················· | ···················· | ···················· | ···················· | ···················· |
| X | 5076 hCoV-19/Netherlands/NB-EMC-384/2020\|EPI_ISL_722585 | ···················· | ···················· | ···················· | ···················· | ···················· | ···················· |
| X | 5077 hCoV-19/Netherlands/UT-EMC-161/2020\|EPI_ISL_722586 | ···················· | ···················· | ···················· | ···················· | ···················· | ···················· |
| X | 5078 hCoV-19/Netherlands/UT-EMC-163/2020\|EPI_ISL_722587 | ···················· | ···················· | ···················· | ···················· | ···················· | ···················· |
| X | 5079 hCoV-19/Netherlands/UT-EMC-164/2020\|EPI_ISL_722588 | ···················· | ···················· | ···················· | ···················· | ···················· | ···················· |
| X | 5080 hCoV-19/Netherlands/ZH-EMC-1079/2020\|EPI_ISL_72258 | ···················· | ···················· | ···················· | ···················· | ···················· | ···················· |
| X | 5081 hCoV-19/Netherlands/ZH-EMC-965/2020\|EPI_ISL_722590 | ···················· | ···················· | ···················· | ···················· | ···················· | ···················· |
| X | 5082 hCoV-19/Netherlands/NB-EMC-355/2020\|EPI_ISL_722591 | ···················· | ···················· | ···················· | ···················· | ···················· | ···················· |
| X | 5083 hCoV-19/Netherlands/NB-EMC-387/2020\|EPI_ISL_722592 | ···················· | ···················· | ···················· | ···················· | ···················· | ···················· |
| X | 5084 hCoV-19/Netherlands/ZH-EMC-1069/2020\|EPI_ISL_72259 | ···················· | ···················· | ···················· | ···················· | ···················· | ···················· |
| X | 5085 hCoV-19/Netherlands/ZH-EMC-952/2020\|EPI_ISL_722594 | ···················· | ···················· | ···················· | ···················· | ······T············· | ···················· |
| X | 5086 hCoV-19/Netherlands/ZH-EMC-972/2020\|EPI_ISL_722595 | ···················· | ···················· | ···················· | ···················· | ···················· | ···················· |
| X | 5087 hCoV-19/Netherlands/ZH-EMC-941/2020\|EPI_ISL_722596 | ···················· | ···················· | ···················· | ···················· | ···················· | ···················· |
| X | 5088 hCoV-19/Netherlands/GE-EMC-358/2020\|EPI_ISL_722597 | ···················· | ···················· | ···················· | ···················· | ···················· | ···················· |
| X | 5089 hCoV-19/Netherlands/GE-EMC-370/2020\|EPI_ISL_722598 | ···················· | ···················· | ···················· | ···················· | ···················· | ···················· |
| X | 5090 hCoV-19/Netherlands/GE-EMC-371/2020\|EPI_ISL_722599 | ···················· | ···················· | ···················· | ···················· | ···················· | ···················· |
| X | 5091 hCoV-19/Netherlands/LI-EMC-109/2020\|EPI_ISL_722600 | ············T······· | ···················· | ···················· | ···················· | ···················· | ···················· |
| X | 5092 hCoV-19/Netherlands/un-EMC-797/2020\|EPI_ISL_722601 | ···················· | ···················· | ···················· | ···················· | ···················· | ···················· |
| X | 5093 hCoV-19/Netherlands/UT-EMC-158/2020\|EPI_ISL_722602 | ···················· | ···················· | ···················· | ···················· | ···················· | ···················· |
| X | 5094 hCoV-19/England/CAMB-1BC20C/2020\|EPI_ISL_723857\|20 | ···················· | ···················· | ···················· | ···················· | ···················· | ···················· |
| X | 5095 hCoV-19/England/CAMB-1BC248/2020\|EPI_ISL_723858\|20 | ···················· | ···················· | ···················· | ···················· | ···················· | ···················· |
| X | 5096 hCoV-19/England/CAMB-1BC3FA/2020\|EPI_ISL_723870\|20 | ···················· | ···················· | ···················· | ···················· | ···················· | ···················· |
| X | 5097 hCoV-19/England/CAMB-1BC424/2020\|EPI_ISL_723871\|20 | ···················· | ···················· | ···················· | ···················· | ···················· | ···················· |
| X | 5098 hCoV-19/England/CAMB-1BC433/2020\|EPI_ISL_723872\|20 | ···················· | ···················· | ···················· | ···················· | ···················· | ···················· |
| X | 5099 hCoV-19/England/CAMB-1BC460/2020\|EPI_ISL_723874\|20 | ···················· | ···················· | ···················· | ···················· | ···················· | ···················· |
| X | 5100 hCoV-19/England/CAMB-1BC48E/2020\|EPI_ISL_723876\|20 | ···················· | ···················· | ···················· | ···················· | ···················· | ···················· |
| X | 5101 hCoV-19/England/CAMB-1BC4AC/2020\|EPI_ISL_723877\|20 | ···················· | ···················· | ···················· | ···················· | ···················· | ···················· |
| X | 5102 hCoV-19/England/CAMB-1BCE08/2020\|EPI_ISL_723900\|20 | ···················· | ···················· | ···················· | ···················· | ···················· | ···················· |
| X | 5103 hCoV-19/England/CAMB-1BCEAE/2020\|EPI_ISL_723905\|20 | ···················· | ·A·················· | ···················· | ···················· | ···················· | ···A················ |
| X | 5104 hCoV-19/England/CAMB-1BCECC/2020\|EPI_ISL_723906\|20 | ···················· | ·A·················· | ···················· | ···················· | ···················· | ···A················ |
| X | 5105 hCoV-19/England/CAMB-1BCEDB/2020\|EPI_ISL_723907\|20 | ···················· | ·A·················· | ···················· | ···················· | ···················· | ···A················ |
| X | 5106 hCoV-19/England/CAMB-1BCEF9/2020\|EPI_ISL_723908\|20 | ···················· | ···················· | ···················· | ···················· | ···················· | ···················· |
| X | 5107 hCoV-19/England/CAMB-1BCF14/2020\|EPI_ISL_723910\|20 | ···················· | ···················· | ···················· | ···················· | ······T············· | ···················· |
| X | 5108 hCoV-19/England/CAMB-1BCF23/2020\|EPI_ISL_723911\|20 | ···················· | ···················· | ···················· | ···················· | ···················· | ···················· |
| X | 5109 hCoV-19/England/CAMB-1BCF32/2020\|EPI_ISL_723912\|20 | ···················· | ···················· | ···················· | ···················· | ···················· | ···················· |
| X | 5110 hCoV-19/England/CAMB-1BCFBA/2020\|EPI_ISL_723917\|20 | ···················· | ···················· | ···················· | ···················· | ···················· | ···················· |
| X | 5111 hCoV-19/England/CAMB-1BCFC9/2020\|EPI_ISL_723918\|20 | ···················· | ···················· | ···················· | ···················· | ···················· | ···················· |
| X | 5112 hCoV-19/England/CAMB-1BCFF6/2020\|EPI_ISL_723920\|20 | ···················· | ···················· | ···················· | ···················· | ···················· | ···················· |
| X | 5113 hCoV-19/England/CAMB-1BD001/2020\|EPI_ISL_723921\|20 | ···················· | ···················· | ···················· | ···················· | ···················· | ···················· |
| X | 5114 hCoV-19/England/CAMB-A977A5/2020\|EPI_ISL_723922\|20 | ···················· | ···················· | ···················· | ···················· | ···················· | ···················· |
| X | 5115 hCoV-19/England/CAMB-A977F0/2020\|EPI_ISL_723924\|20 | ···················· | ···················· | ···················· | ···················· | ···················· | ···················· |
| X | 5116 hCoV-19/England/CAMB-A97848/2020\|EPI_ISL_723927\|20 | ···················· | ···················· | ···················· | ···················· | ···················· | ···················· |
| X | 5117 hCoV-19/England/CAMB-A97866/2020\|EPI_ISL_723928\|20 | ···················· | ···················· | ···················· | ···················· | ···················· | ···················· |
| X | 5118 hCoV-19/England/CAMB-A97893/2020\|EPI_ISL_723930\|20 | ···················· | ···················· | ···················· | ···················· | ···················· | ···················· |
| X | 5119 hCoV-19/England/CAMB-A978A2/2020\|EPI_ISL_723931\|20 | ···················· | ···················· | ···················· | ···················· | ···················· | ···················· |
| X | 5120 hCoV-19/England/CAMB-A978C0/2020\|EPI_ISL_723932\|20 | ···················· | ···················· | ···················· | ···················· | ···················· | ···················· |
| X | 5121 hCoV-19/Wales/ALDP-BE9178/2020\|EPI_ISL_710625\|2020 | ···················· | ···················· | ···················· | ···················· | ···················· | ···················· |
| X | 5122 hCoV-19/Scotland/QEUH-BEB185/2020\|EPI_ISL_710627\|2 | ···················· | ···················· | ···················· | ···················· | ···················· | ···················· |
| X | 5123 hCoV-19/Wales/ALDP-BE9196/2020\|EPI_ISL_710628\|2020 | ···················· | ···················· | ···················· | ···················· | ···················· | ···················· |
| X | 5124 hCoV-19/Scotland/QEUH-BEB1FE/2020\|EPI_ISL_710629\|2 | ···················· | ···················· | ···················· | ···················· | ···················· | ···················· |
| X | 5125 hCoV-19/Wales/ALDP-BE8E90/2020\|EPI_ISL_710631\|2020 | ···················· | ···················· | ···················· | ···················· | ···················· | ···················· |
| X | 5126 hCoV-19/Scotland/QEUH-BEAF22/2020\|EPI_ISL_710633\|2 | ···················· | ·A·················· | ···················· | ···················· | ···················· | ···A················ |
| X | 5127 hCoV-19/Scotland/QEUH-BEB194/2020\|EPI_ISL_710635\|2 | ···················· | ···················· | ···················· | ···················· | ···················· | ···················· |
| X | 5128 hCoV-19/Scotland/QEUH-BEB1C1/2020\|EPI_ISL_710637\|2 | ···················· | ···················· | ···················· | ···················· | ···················· | ···················· |
| X | 5129 hCoV-19/Wales/ALDP-BE8E18/2020\|EPI_ISL_710638\|2020 | ···················· | ···················· | ···················· | ···················· | ···················· | ···················· |
| X | 5130 hCoV-19/Scotland/QEUH-BEB167/2020\|EPI_ISL_710640\|2 | ···················· | ···················· | ···················· | ···················· | ···················· | ···················· |
| X | 5131 hCoV-19/Scotland/QEUH-BEAE8F/2020\|EPI_ISL_710641\|2 | ···················· | ···················· | ···················· | ···················· | ···················· | ···················· |
| X | 5132 hCoV-19/Scotland/QEUH-BEB1EF/2020\|EPI_ISL_710642\|2 | ···················· | ···················· | ···················· | ···················· | ···················· | ···················· |
| X | 5133 hCoV-19/Scotland/QEUH-BEAF5F/2020\|EPI_ISL_710643\|2 | ···················· | ···················· | ···················· | ···················· | ···················· | ···················· |
| X | 5134 hCoV-19/Scotland/QEUH-BEAF31/2020\|EPI_ISL_710644\|2 | ···················· | ···················· | ···················· | ···················· | ···················· | ···················· |
| X | 5135 hCoV-19/Wales/ALDP-BE961F/2020\|EPI_ISL_710645\|2020 | ···················· | ···················· | ···················· | ···················· | ···················· | ···················· |
| X | 5136 hCoV-19/Scotland/QEUH-BEB04C/2020\|EPI_ISL_710646\|2 | ···················· | ···················· | ···················· | ···················· | ···················· | ···················· |
| X | 5137 hCoV-19/Wales/ALDP-BE8E54/2020\|EPI_ISL_710647\|2020 | ···················· | ···················· | ···················· | ···················· | ···················· | ···················· |
| X | 5139 hCoV-19/Scotland/QEUH-BF19A2/2020\|EPI_ISL_710651\|2 | ···················· | ···················· | ···················· | ···················· | ···················· | ···················· |
| X | 5140 hCoV-19/Scotland/QEUH-BF1993/2020\|EPI_ISL_710652\|2 | ···················· | ···················· | ···················· | ···················· | ···················· | ···················· |

| | | | | | | | | |
|---|---|---|---|---|---|---|---|---|
| X | 5141 | hCoV-19/Scotland/QEUH-C1488D/2020\|EPI_ISL_710654\|2 | ···················· | ···················· | ···················· | ···················· | ···················· | ···················· |
| X | 5142 | hCoV-19/Scotland/QEUH-C14C72/2020\|EPI_ISL_710655\|2 | ···················· | ···················· | ···················· | ···················· | ···················· | ···················· |
| X | 5143 | hCoV-19/Wales/ALDP-C062BD/2020\|EPI_ISL_710656\|2020 | ···················· | ···················· | ···················· | ···················· | ···················· | ···················· |
| X | 5144 | hCoV-19/Scotland/QEUH-C148D8/2020\|EPI_ISL_710657\|2 | ···················· | ···················· | ···················· | ···················· | ···················· | ···················· |
| X | 5145 | hCoV-19/Scotland/QEUH-C14CEB/2020\|EPI_ISL_710658\|2 | ···················· | ···················· | ···················· | ···················· | ···················· | ···················· |
| X | 5146 | hCoV-19/Scotland/QEUH-C14C90/2020\|EPI_ISL_710659\|2 | ···················· | ···················· | ···················· | ···················· | ···················· | ···················· |
| X | 5147 | hCoV-19/Scotland/QEUH-C14CCD/2020\|EPI_ISL_710660\|2 | ···················· | ···················· | ···················· | ···················· | ···················· | ···················· |
| X | 5148 | hCoV-19/Wales/ALDP-C067FA/2020\|EPI_ISL_710661\|2020 | ···················· | ···················· | ···················· | ···················· | ···················· | ···················· |
| X | 5149 | hCoV-19/Wales/ALDP-C068E8/2020\|EPI_ISL_710662\|2020 | ···················· | ···················· | ···················· | ···················· | ······T············· | ···················· |
| X | 5150 | hCoV-19/Wales/ALDP-C06578/2020\|EPI_ISL_710663\|2020 | ···················· | ···················· | ···················· | ···················· | ···················· | ···················· |
| X | 5151 | hCoV-19/Scotland/QEUH-C1691F/2020\|EPI_ISL_710664\|2 | ···················· | ···················· | ···················· | ···················· | ···················· | ···················· |
| X | 5152 | hCoV-19/Wales/ALDP-C0651E/2020\|EPI_ISL_710665\|2020 | ···················· | ···················· | ···················· | ···················· | ···················· | ···················· |
| X | 5153 | hCoV-19/Scotland/QEUH-C1486F/2020\|EPI_ISL_710666\|2 | ···················· | ···················· | ···················· | ···················· | ···················· | ···················· |
| X | 5154 | hCoV-19/Scotland/QEUH-C14CBE/2020\|EPI_ISL_710667\|2 | ···················· | ···················· | ···················· | ···················· | ···················· | ···················· |
| X | 5155 | hCoV-19/Scotland/QEUH-C1694C/2020\|EPI_ISL_710668\|2 | ···················· | ···················· | ···················· | ···················· | ···················· | ···················· |
| X | 5156 | hCoV-19/Scotland/QEUH-C14902/2020\|EPI_ISL_710669\|2 | ···················· | ···················· | ···················· | ···················· | ···················· | ···················· |
| X | 5157 | hCoV-19/Scotland/QEUH-C14920/2020\|EPI_ISL_710675\|2 | ···················· | ···················· | ···················· | ···················· | ···················· | ···················· |
| X | 5158 | hCoV-19/Wales/ALDP-C0694F/2020\|EPI_ISL_710676\|2020 | ···················· | ···················· | ···················· | ···················· | ···················· | ···················· |
| X | 5159 | hCoV-19/Scotland/QEUH-C14CDC/2020\|EPI_ISL_710677\|2 | ···················· | ···················· | ···················· | ···················· | ···················· | ···················· |
| X | 5160 | hCoV-19/Scotland/QEUH-C14CFA/2020\|EPI_ISL_710678\|2 | ···················· | ···················· | ···················· | ···················· | ···················· | ···················· |
| X | 5161 | hCoV-19/Scotland/QEUH-C1696A/2020\|EPI_ISL_710690\|2 | ···················· | ···················· | ···················· | ···················· | ···················· | ···················· |
| X | 5162 | hCoV-19/Scotland/QEUH-C14C63/2020\|EPI_ISL_710691\|2 | ···················· | ···················· | ···················· | ···················· | ···················· | ···················· |
| X | 5163 | hCoV-19/Wales/ALDP-C06569/2020\|EPI_ISL_710693\|2020 | ···················· | ···················· | ···················· | ···················· | ···················· | ···················· |
| X | 5164 | hCoV-19/Wales/ALDP-C06842/2020\|EPI_ISL_710694\|2020 | ···················· | ···················· | ···················· | ···················· | ···················· | ···················· |
| X | 5165 | hCoV-19/Scotland/QEUH-C148BA/2020\|EPI_ISL_710696\|2 | ···················· | ···················· | ···················· | ···················· | ···················· | ···················· |
| X | 5166 | hCoV-19/Scotland/QEUH-C14CAF/2020\|EPI_ISL_710697\|2 | ···················· | ···················· | ···················· | ···················· | ···················· | ···················· |
| X | 5167 | hCoV-19/Wales/ALDP-C0639C/2020\|EPI_ISL_710698\|2020 | ···················· | ···················· | ···················· | ···················· | ···················· | ···················· |
| X | 5168 | hCoV-19/Wales/ALDP-BB5500/2020\|EPI_ISL_710699\|2020 | ···················· | ···················· | ···················· | ···················· | ···················· | ···················· |
| X | 5169 | hCoV-19/Denmark/DCGC-14750/2020\|EPI_ISL_711955\|202 | ···················· | ···················· | ···················· | ···················· | ···················· | ···················· |
| X | 5170 | hCoV-19/Denmark/DCGC-14751/2020\|EPI_ISL_711956\|202 | ···················· | ···················· | ···················· | ···················· | ···················· | ···················· |
| X | 5171 | hCoV-19/Denmark/DCGC-14754/2020\|EPI_ISL_711957\|202 | ···················· | ···················· | ···················· | ···················· | ···················· | ···················· |
| X | 5172 | hCoV-19/Denmark/DCGC-14755/2020\|EPI_ISL_711958\|202 | ···················· | ···················· | ···················· | ···················· | ···················· | ···················· |
| X | 5173 | hCoV-19/Denmark/DCGC-14757/2020\|EPI_ISL_711959\|202 | ···················· | ···················· | ···················· | ···················· | ···················· | ···················· |
| X | 5174 | hCoV-19/Denmark/DCGC-14759/2020\|EPI_ISL_711960\|202 | ···················· | ·A·················· | ···················· | ···················· | ···················· | ···A················ |
| X | 5175 | hCoV-19/Denmark/DCGC-14762/2020\|EPI_ISL_711961\|202 | ···················· | ···················· | ···················· | ···················· | ···················· | ···················· |
| X | 5176 | hCoV-19/Denmark/DCGC-14765/2020\|EPI_ISL_711962\|202 | ···················· | ···················· | ···················· | ···················· | ···················· | ···················· |
| X | 5177 | hCoV-19/Denmark/DCGC-14766/2020\|EPI_ISL_711963\|202 | ···················· | ···················· | ···················· | ···················· | ···················· | ···················· |
| X | 5178 | hCoV-19/Denmark/DCGC-14767/2020\|EPI_ISL_711964\|202 | ···················· | ···················· | ···················· | ···················· | ···················· | ···················· |
| X | 5179 | hCoV-19/Denmark/DCGC-14768/2020\|EPI_ISL_711965\|202 | ···················· | ···················· | ···················· | ···················· | ···················· | ···················· |
| X | 5180 | hCoV-19/Denmark/DCGC-14769/2020\|EPI_ISL_711966\|202 | ···················· | ···················· | ···················· | ···················· | ···················· | ···················· |
| X | 5181 | hCoV-19/Denmark/DCGC-14770/2020\|EPI_ISL_711967\|202 | ···················· | ···················· | ···················· | ···················· | ···················· | ···················· |
| X | 5182 | hCoV-19/Denmark/DCGC-14771/2020\|EPI_ISL_711968\|202 | ···················· | ···················· | ···················· | ···················· | ···················· | ···················· |
| X | 5183 | hCoV-19/Denmark/DCGC-14772/2020\|EPI_ISL_711969\|202 | ···················· | ···················· | ···················· | ···················· | ···················· | ···················· |
| X | 5184 | hCoV-19/Denmark/DCGC-14774/2020\|EPI_ISL_711970\|202 | ···················· | ···················· | ···················· | ···················· | ···················· | ···················· |
| X | 5185 | hCoV-19/Denmark/DCGC-14775/2020\|EPI_ISL_711971\|202 | ···················· | ···················· | ···················· | ···················· | ···················· | ···················· |
| X | 5186 | hCoV-19/Denmark/DCGC-14776/2020\|EPI_ISL_711972\|202 | ···················· | ···················· | ···················· | ···················· | ···················· | ···················· |
| X | 5187 | hCoV-19/Denmark/DCGC-14777/2020\|EPI_ISL_711973\|202 | ···················· | ···················· | ···················· | ···················· | ···················· | ···················· |
| X | 5188 | hCoV-19/Denmark/DCGC-14779/2020\|EPI_ISL_711974\|202 | ···················· | ···················· | ···················· | ···················· | ···················· | ···················· |
| X | 5189 | hCoV-19/Denmark/DCGC-14780/2020\|EPI_ISL_711975\|202 | ···················· | ·A·················· | ···················· | ···················· | ···················· | ···A················ |
| X | 5190 | hCoV-19/Denmark/DCGC-14781/2020\|EPI_ISL_711976\|202 | ···················· | ···················· | ···················· | ···················· | ···················· | ···················· |
| X | 5191 | hCoV-19/Denmark/DCGC-14782/2020\|EPI_ISL_711977\|202 | ···················· | ···················· | ···················· | ···················· | ···················· | ···················· |
| X | 5192 | hCoV-19/Denmark/DCGC-14784/2020\|EPI_ISL_711978\|202 | ···················· | ···················· | ···················· | ···················· | ···················· | ···················· |
| X | 5193 | hCoV-19/Denmark/DCGC-14785/2020\|EPI_ISL_711979\|202 | ···················· | ···················· | ···················· | ···················· | ···················· | ···················· |
| X | 5194 | hCoV-19/Denmark/DCGC-14786/2020\|EPI_ISL_711980\|202 | ···················· | ···················· | ···················· | ···················· | ···················· | ···················· |
| X | 5195 | hCoV-19/Denmark/DCGC-14787/2020\|EPI_ISL_711981\|202 | ···················· | ···················· | ···················· | ···················· | ···················· | ···················· |
| X | 5196 | hCoV-19/Denmark/DCGC-14789/2020\|EPI_ISL_711982\|202 | ···················· | ···················· | ···················· | ···················· | ···················· | ···················· |
| X | 5197 | hCoV-19/Denmark/DCGC-14790/2020\|EPI_ISL_711983\|202 | ···················· | ···················· | ···················· | ···················· | ···················· | ···················· |
| X | 5198 | hCoV-19/Denmark/DCGC-14791/2020\|EPI_ISL_711984\|202 | ···················· | ···················· | ···················· | ···················· | ···················· | ···················· |
| X | 5199 | hCoV-19/Denmark/DCGC-14792/2020\|EPI_ISL_711985\|202 | ···················· | ···················· | ···················· | ···················· | ···················· | ···················· |
| X | 5200 | hCoV-19/Denmark/DCGC-14793/2020\|EPI_ISL_711986\|202 | ···················· | ···················· | ···················· | ···················· | ···················· | ···················· |
| X | 5201 | hCoV-19/Denmark/DCGC-14794/2020\|EPI_ISL_711987\|202 | ···················· | ···················· | ···················· | ···················· | ···················· | ···················· |
| X | 5202 | hCoV-19/Denmark/DCGC-14795/2020\|EPI_ISL_711988\|202 | ···················· | ·A·················· | ···················· | ···················· | ···················· | ···················· |
| X | 5203 | hCoV-19/Denmark/DCGC-14797/2020\|EPI_ISL_711989\|202 | ···················· | ···················· | ···················· | ···················· | ···················· | ···················· |
| X | 5204 | hCoV-19/Denmark/DCGC-14799/2020\|EPI_ISL_711990\|202 | ···················· | ···················· | ···················· | ···················· | ···················· | ···················· |
| X | 5205 | hCoV-19/Denmark/DCGC-14800/2020\|EPI_ISL_711991\|202 | ···················· | ···················· | ···················· | ···················· | ···················· | ···················· |
| X | 5206 | hCoV-19/Denmark/DCGC-14801/2020\|EPI_ISL_711992\|202 | ···················· | ···················· | ···················· | ···················· | ···················· | ···················· |
| X | 5207 | hCoV-19/Denmark/DCGC-14802/2020\|EPI_ISL_711993\|202 | ···················· | ···················· | ···················· | ···················· | ···················· | ···················· |
| X | 5208 | hCoV-19/Denmark/DCGC-14803/2020\|EPI_ISL_711994\|202 | ···················· | ···················· | ···················· | ···················· | ···················· | ···················· |
| X | 5209 | hCoV-19/Denmark/DCGC-14804/2020\|EPI_ISL_711995\|202 | ···················· | ·A·················· | ···················· | ···················· | ···················· | ···A················ |
| X | 5210 | hCoV-19/Denmark/DCGC-14805/2020\|EPI_ISL_711996\|202 | ···················· | ···················· | ···················· | ···················· | ···················· | ···················· |
| X | 5211 | hCoV-19/Denmark/DCGC-14806/2020\|EPI_ISL_711997\|202 | ···················· | ···················· | ···················· | ···················· | ···················· | ···················· |
| X | 5212 | hCoV-19/Denmark/DCGC-14807/2020\|EPI_ISL_711998\|202 | ···················· | ···················· | ···················· | ···················· | ···················· | ···················· |
| X | 5213 | hCoV-19/Denmark/DCGC-14808/2020\|EPI_ISL_711999\|202 | ···················· | ···················· | ···················· | ···················· | ···················· | ···················· |
| X | 5214 | hCoV-19/England/CAMC-C3E638/2020\|EPI_ISL_719001\|20 | ···················· | ···················· | ···················· | ···················· | ···················· | ···················· |
| X | 5215 | hCoV-19/England/CAMC-C41BD9/2020\|EPI_ISL_719002\|20 | ···················· | ···················· | ···················· | ···················· | ···················· | ···················· |
| X | 5216 | hCoV-19/England/CAMC-C3EE4F/2020\|EPI_ISL_719006\|20 | ···················· | ···················· | ···················· | ···················· | ···················· | ···················· |
| X | 5217 | hCoV-19/England/CAMC-C3EDE8/2020\|EPI_ISL_719008\|20 | ···················· | ···················· | ···················· | ···················· | ···················· | ···················· |
| X | 5218 | hCoV-19/England/CAMC-C438B2/2020\|EPI_ISL_719022\|20 | ···················· | ···················· | ···················· | ···················· | ···················· | ···················· |
| X | 5219 | hCoV-19/England/CAMC-C438A3/2020\|EPI_ISL_719030\|20 | ···················· | ···················· | ···················· | ···················· | ···················· | ···················· |
| X | 5220 | hCoV-19/England/CAMC-C4412F/2020\|EPI_ISL_719032\|20 | ···················· | ···················· | ···················· | ···················· | ···················· | ···················· |

| | | | | | | | | |
|---|---|---|---|---|---|---|---|---|
| X | 5221 | hCoV-19/England/MILK-BB1373/2020\|EPI_ISL_720008\|20 | ···················· | ···················· | ···················· | ···················· | ···················· | ···················· |
| X | 5222 | hCoV-19/England/MILK-BB174A/2020\|EPI_ISL_720009\|20 | ···················· | ···················· | ···················· | ···················· | ···················· | ···················· |
| X | 5223 | hCoV-19/England/MILK-BB1391/2020\|EPI_ISL_720011\|20 | ···················· | ···················· | ···················· | ···················· | ···················· | ···················· |
| X | 5224 | hCoV-19/England/MILK-BB20E4/2020\|EPI_ISL_720013\|20 | ···················· | ···················· | ···················· | ···················· | ···················· | ···················· |
| X | 5225 | hCoV-19/England/MILK-BB08A2/2020\|EPI_ISL_720017\|20 | ···················· | ···················· | ···················· | ···················· | ···················· | ···················· |
| X | 5226 | hCoV-19/England/MILK-BB0A8E/2020\|EPI_ISL_720019\|20 | ···················· | ···················· | ···················· | ···················· | ···················· | ···················· |
| X | 5227 | hCoV-19/England/MILK-BB1443/2020\|EPI_ISL_720020\|20 | ···················· | ···················· | ···················· | ···················· | ···················· | ···················· |
| X | 5228 | hCoV-19/England/MILK-BB199F/2020\|EPI_ISL_720021\|20 | ···················· | ···················· | ···················· | ···················· | ···················· | ···················· |
| X | 5229 | hCoV-19/England/MILK-BB016B/2020\|EPI_ISL_720023\|20 | ···················· | ···················· | ···················· | ···················· | ···················· | ···················· |
| X | 5230 | hCoV-19/England/MILK-BB1425/2020\|EPI_ISL_720024\|20 | ···················· | ···················· | ···················· | ···················· | ···················· | ···················· |
| X | 5231 | hCoV-19/England/MILK-BB13CE/2020\|EPI_ISL_720025\|20 | ···················· | ···················· | ···················· | ···················· | ···················· | ···················· |
| X | 5232 | hCoV-19/England/ALDP-BC7A93/2020\|EPI_ISL_721300\|20 | ···················· | ···················· | ···················· | ···················· | ···················· | ···················· |
| X | 5233 | hCoV-19/England/ALDP-BC7A1B/2020\|EPI_ISL_721301\|20 | ···················· | ···················· | ···················· | ···················· | ···················· | ···················· |
| X | 5234 | hCoV-19/England/ALDP-BC8713/2020\|EPI_ISL_721302\|20 | ···················· | ···················· | ···················· | ···················· | ···················· | ···················· |
| X | 5235 | hCoV-19/England/ALDP-BC8889/2020\|EPI_ISL_721303\|20 | ···················· | ···················· | ···················· | ···················· | ···················· | ···················· |
| X | 5236 | hCoV-19/England/ALDP-BC85FB/2020\|EPI_ISL_721305\|20 | ···················· | ···················· | ···················· | ···················· | ···················· | ···················· |
| X | 5237 | hCoV-19/England/ALDP-BC64D2/2020\|EPI_ISL_721306\|20 | ···················· | ···················· | ···················· | ···················· | ···················· | ···················· |
| X | 5238 | hCoV-19/England/ALDP-BC65B1/2020\|EPI_ISL_721307\|20 | ···················· | ···················· | ···················· | ···················· | ···················· | ···················· |
| X | 5239 | hCoV-19/England/ALDP-BC74FF/2020\|EPI_ISL_721308\|20 | ···················· | ···················· | ···················· | ···················· | ···················· | ···················· |
| X | 5240 | hCoV-19/England/ALDP-BC627E/2020\|EPI_ISL_721309\|20 | ···················· | ···················· | ···················· | ···················· | ···················· | ···A················ |
| X | 5241 | hCoV-19/England/ALDP-BC77D8/2020\|EPI_ISL_721310\|20 | ···················· | ···················· | ···················· | ···················· | ···················· | ···················· |
| X | 5242 | hCoV-19/England/ALDP-BC743B/2020\|EPI_ISL_721311\|20 | ···················· | ···················· | ···················· | ···················· | ···················· | ···················· |
| X | 5243 | hCoV-19/England/ALDP-BC8731/2020\|EPI_ISL_721312\|20 | ···················· | ···················· | ···················· | ···················· | ···················· | ···················· |
| X | 5244 | hCoV-19/England/ALDP-BC791E/2020\|EPI_ISL_721313\|20 | ···················· | ···················· | ···················· | ···················· | ···················· | ···················· |
| X | 5245 | hCoV-19/England/ALDP-BC795A/2020\|EPI_ISL_721314\|20 | ···················· | ···················· | ···················· | ···················· | ···················· | ···················· |
| X | 5246 | hCoV-19/England/ALDP-BC7B27/2020\|EPI_ISL_721322\|20 | ···················· | ···················· | ···················· | ···················· | ···················· | ···················· |
| X | 5247 | hCoV-19/England/ALDP-BC7CCA/2020\|EPI_ISL_721323\|20 | ···················· | ···················· | ···················· | ···················· | ···················· | ···················· |
| X | 5248 | hCoV-19/England/ALDP-BC6171/2020\|EPI_ISL_721324\|20 | ···················· | ···················· | ···················· | ···················· | ······T············· | ···················· |
| X | 5249 | hCoV-19/England/ALDP-BC67BB/2020\|EPI_ISL_721326\|20 | ···················· | ···················· | ···················· | ···················· | ···················· | ···················· |
| X | 5250 | hCoV-19/England/ALDP-BC61DB/2020\|EPI_ISL_721338\|20 | ···················· | ···················· | ···················· | ···················· | ···················· | ···················· |
| X | 5251 | hCoV-19/England/ALDP-BC63D5/2020\|EPI_ISL_721339\|20 | ···················· | ···················· | ···················· | ···················· | ···················· | ···················· |
| X | 5252 | hCoV-19/England/ALDP-BC741D/2020\|EPI_ISL_721340\|20 | ···················· | ···················· | ···················· | ···················· | ···················· | ···················· |
| X | 5253 | hCoV-19/England/ALDP-BC6812/2020\|EPI_ISL_721342\|20 | ···················· | ···················· | ···················· | ···················· | ···················· | ···················· |
| X | 5254 | hCoV-19/England/ALDP-BC75A1/2020\|EPI_ISL_721343\|20 | ···················· | ···················· | ···················· | ···················· | ···················· | ···················· |
| X | 5255 | hCoV-19/England/ALDP-BC6663/2020\|EPI_ISL_721344\|20 | ···················· | ···················· | ···················· | ···················· | ···················· | ···················· |
| X | 5256 | hCoV-19/England/ALDP-BC7556/2020\|EPI_ISL_721346\|20 | ···················· | ···················· | ···················· | ···················· | ···················· | ···················· |
| X | 5257 | hCoV-19/England/ALDP-BC794B/2020\|EPI_ISL_721347\|20 | ···················· | ···················· | ···················· | ···················· | ···················· | ···················· |
| X | 5258 | hCoV-19/England/ALDP-BC7C15/2020\|EPI_ISL_721348\|20 | ···················· | ···················· | ···················· | ···················· | ···················· | ···················· |
| X | 5259 | hCoV-19/England/ALDP-BC7CAC/2020\|EPI_ISL_721349\|20 | ···················· | ···················· | ···················· | ···················· | ···················· | ···················· |
| X | 5260 | hCoV-19/England/ALDP-BC7B36/2020\|EPI_ISL_721350\|20 | ···················· | ···················· | ···················· | ···················· | ···················· | ···················· |
| X | 5261 | hCoV-19/England/ALDP-BC73B6/2020\|EPI_ISL_721352\|20 | ···················· | ···················· | ···················· | ···················· | ···················· | ···················· |
| X | 5262 | hCoV-19/England/ALDP-BC8564/2020\|EPI_ISL_721355\|20 | ···················· | ···················· | ···················· | ···················· | ···················· | ···················· |
| X | 5263 | hCoV-19/England/ALDP-BC7B90/2020\|EPI_ISL_721357\|20 | ···················· | ···················· | ···················· | ···················· | ···················· | ···················· |
| X | 5264 | hCoV-19/England/ALDP-BC636C/2020\|EPI_ISL_721358\|20 | ···················· | ···················· | ···················· | ···················· | ······T············· | ···················· |
| X | 5265 | hCoV-19/England/ALDP-BC78B7/2020\|EPI_ISL_721359\|20 | ···················· | ···················· | ···················· | ···················· | ···················· | ···················· |
| X | 5266 | hCoV-19/Netherlands/UT-EMC-165/2020\|EPI_ISL_722603 | ···················· | ···················· | ···················· | ···················· | ···················· | ···················· |
| X | 5267 | hCoV-19/Netherlands/UT-EMC-166/2020\|EPI_ISL_722604 | ···················· | ···················· | ···················· | ···················· | ···················· | ···················· |
| X | 5268 | hCoV-19/Netherlands/ZH-EMC-1011/2020\|EPI_ISL_72260 | ···················· | ···················· | ···················· | ···················· | ···················· | ···················· |
| X | 5269 | hCoV-19/Netherlands/ZH-EMC-1031/2020\|EPI_ISL_72260 | ···················· | ···················· | ···················· | ···················· | ······T············· | ···················· |
| X | 5270 | hCoV-19/Netherlands/ZH-EMC-966/2020\|EPI_ISL_722607 | ···················· | ···················· | ···················· | ···················· | ···················· | ···················· |
| X | 5271 | hCoV-19/Netherlands/GE-EMC-375/2020\|EPI_ISL_722608 | ···················· | ···················· | ···················· | ···················· | ···················· | ···················· |
| X | 5272 | hCoV-19/Netherlands/NB-EMC-419/2020\|EPI_ISL_722609 | ···················· | ···················· | ···················· | ···················· | ···················· | ···················· |
| X | 5273 | hCoV-19/Netherlands/NB-EMC-425/2020\|EPI_ISL_722610 | ···················· | ···················· | ···················· | ···················· | ···················· | ···················· |
| X | 5274 | hCoV-19/Netherlands/UT-EMC-207/2020\|EPI_ISL_722611 | ···················· | ···················· | ···················· | ···················· | ···················· | ···················· |
| X | 5275 | hCoV-19/Netherlands/FL-EMC-44/2020\|EPI_ISL_722612\| | ···················· | ···················· | ···················· | ···················· | ···················· | ···················· |
| X | 5276 | hCoV-19/Netherlands/GE-EMC-354/2020\|EPI_ISL_722613 | ···················· | ···················· | ···················· | ···················· | ···················· | ···················· |
| X | 5277 | hCoV-19/Netherlands/NB-EMC-420/2020\|EPI_ISL_722614 | ···················· | ···················· | ···················· | ···················· | ···················· | ···················· |
| X | 5278 | hCoV-19/Netherlands/NB-EMC-423/2020\|EPI_ISL_722615 | ···················· | ···················· | ···················· | ···················· | ···················· | ···················· |
| X | 5279 | hCoV-19/Netherlands/NB-EMC-424/2020\|EPI_ISL_722616 | ···················· | ···················· | ···················· | ···················· | ···················· | ···················· |
| X | 5280 | hCoV-19/Netherlands/ZH-EMC-1009/2020\|EPI_ISL_72261 | ···················· | ···················· | ···················· | ···················· | ···················· | ···················· |
| X | 5281 | hCoV-19/Netherlands/ZH-EMC-1021/2020\|EPI_ISL_72261 | ···················· | ···················· | ···················· | ···················· | ···················· | ···················· |
| X | 5282 | hCoV-19/Netherlands/GE-EMC-368/2020\|EPI_ISL_722619 | ···················· | ···················· | ···················· | ···················· | ······T············· | ···················· |
| X | 5283 | hCoV-19/Netherlands/GE-EMC-408/2020\|EPI_ISL_722620 | ···················· | ···················· | ···················· | ···················· | ···················· | ···················· |
| X | 5284 | hCoV-19/Netherlands/GE-EMC-410/2020\|EPI_ISL_722621 | ···················· | ···················· | ···················· | ···················· | ···················· | ···················· |
| X | 5285 | hCoV-19/Netherlands/NB-EMC-421/2020\|EPI_ISL_722622 | ···················· | ···················· | ···················· | ···················· | ···················· | ···················· |
| X | 5286 | hCoV-19/Netherlands/NB-EMC-426/2020\|EPI_ISL_722623 | ···················· | ···················· | ···················· | ···················· | ···················· | ···················· |
| X | 5287 | hCoV-19/Netherlands/NB-EMC-429/2020\|EPI_ISL_722624 | ···················· | ···················· | ···················· | ···················· | ···················· | ···················· |
| X | 5288 | hCoV-19/Netherlands/UT-EMC-149/2020\|EPI_ISL_722625 | ···················· | ···················· | ···················· | ···················· | ···················· | ···················· |
| X | 5289 | hCoV-19/Netherlands/ZH-EMC-1016/2020\|EPI_ISL_72262 | ···················· | ···················· | ···················· | ···················· | ···················· | ···················· |
| X | 5290 | hCoV-19/Netherlands/ZH-EMC-1025/2020\|EPI_ISL_72262 | ···················· | ···················· | ···················· | ···················· | ···················· | ···················· |
| X | 5291 | hCoV-19/Netherlands/ZH-EMC-980/2020\|EPI_ISL_722628 | ···················· | ···················· | ···················· | ···················· | ···················· | ···················· |
| X | 5292 | hCoV-19/Netherlands/GE-EMC-367/2020\|EPI_ISL_722629 | ···················· | ···················· | ···················· | ···················· | ···················· | ···················· |
| X | 5293 | hCoV-19/Netherlands/GE-EMC-389/2020\|EPI_ISL_722630 | ···················· | ···················· | ···················· | ···················· | ···················· | ···················· |
| X | 5294 | hCoV-19/Netherlands/UT-EMC-194/2020\|EPI_ISL_722631 | ···················· | ···················· | ···················· | ···················· | ···················· | ···················· |
| X | 5295 | hCoV-19/Netherlands/ZE-EMC-83/2020\|EPI_ISL_722632\| | ···················· | ···················· | ···················· | ···················· | ···················· | ···················· |
| X | 5296 | hCoV-19/Netherlands/ZH-EMC-1005/2020\|EPI_ISL_72263 | ···················· | ···················· | ···················· | ···················· | ···················· | ···················· |
| X | 5297 | hCoV-19/Netherlands/ZH-EMC-1018/2020\|EPI_ISL_72263 | ···················· | ···················· | ···················· | ···················· | ······T············· | ···················· |
| X | 5298 | hCoV-19/Netherlands/ZH-EMC-956/2020\|EPI_ISL_722635 | ···················· | ···················· | ···················· | ···················· | ······T············· | ···················· |
| X | 5299 | hCoV-19/Netherlands/ZH-EMC-993/2020\|EPI_ISL_722636 | ···················· | ···················· | ···················· | ···················· | ······T············· | ···················· |
| X | 5300 | hCoV-19/Netherlands/NB-EMC-436/2020\|EPI_ISL_722637 | ···················· | ···················· | ···················· | ···················· | ···················· | ···················· |

X 5301 hCoV-19/Netherlands/UT-EMC-150/2020|EPI_ISL_722638 ···················· ···················· ····················· ···················· ···················· ····················
X 5302 hCoV-19/Netherlands/UT-EMC-203/2020|EPI_ISL_722639 ···················· ···················· ····················· ···················· ···················· ····················
X 5303 hCoV-19/Netherlands/ZE-EMC-80/2020|EPI_ISL_722640| ···················· ···················· ····················· ···················· ···················· ····················
X 5304 hCoV-19/Netherlands/ZH-EMC-984/2020|EPI_ISL_722641 ···················· ···················· ····················· ···················· ···················· ····················
X 5305 hCoV-19/Netherlands/GE-EMC-350/2020|EPI_ISL_722642 ···················· ···················· ····················· ···················· ······T············· ····················
X 5306 hCoV-19/Netherlands/GE-EMC-414/2020|EPI_ISL_722643 ···················· ···················· ····················· ···················· ···················· ····················
X 5307 hCoV-19/Netherlands/GE-EMC-418/2020|EPI_ISL_722644 ···················· ···················· ····················· ···················· ······T············· ····················
X 5308 hCoV-19/Netherlands/NB-EMC-428/2020|EPI_ISL_722645 ···················· ···················· ····················· ···················· ···················· ····················
X 5309 hCoV-19/Netherlands/UT-EMC-153/2020|EPI_ISL_722646 ···················· ···················· ····················· ···················· ···················· ····················
X 5310 hCoV-19/Netherlands/GE-EMC-391/2020|EPI_ISL_722650 ···················· ···················· ····················· ···················· ···················· ····················
X 5311 hCoV-19/Netherlands/NB-EMC-427/2020|EPI_ISL_722651 ···················· ···················· ····················· ···················· ···················· ····················
X 5312 hCoV-19/Netherlands/UT-EMC-154/2020|EPI_ISL_722652 ···················· ···················· ····················· ···················· ···················· ····················
X 5313 hCoV-19/Netherlands/UT-EMC-155/2020|EPI_ISL_722653 ···················· ···················· ····················· ···················· ···················· ····················
X 5314 hCoV-19/Netherlands/UT-EMC-196/2020|EPI_ISL_722654 ···················· ···················· ····················· ···················· ···················· ····················
X 5315 hCoV-19/Netherlands/ZH-EMC-1007/2020|EPI_ISL_72265 ···················· ···················· ····················· ···················· ···················· ····················
X 5316 hCoV-19/Netherlands/ZH-EMC-1010/2020|EPI_ISL_72265 ···················· ···················· ····················· ···················· ······T············· ····················
X 5317 hCoV-19/Netherlands/ZH-EMC-1019/2020|EPI_ISL_72265 ···················· ···················· ····················· ···················· ······T············· ····················
X 5318 hCoV-19/Netherlands/UT-EMC-198/2020|EPI_ISL_722669 ···················· ···················· ····················· ···················· ···················· ····················
X 5319 hCoV-19/Netherlands/UT-EMC-209/2020|EPI_ISL_722670 ···················· ···················· ····················· ···················· ···················· ····················
X 5320 hCoV-19/Netherlands/ZH-EMC-1003/2020|EPI_ISL_72267 ···················· ···················· ····················· ···················· ···················· ····················
X 5321 hCoV-19/Netherlands/ZH-EMC-1008/2020|EPI_ISL_72267 ···················· ···················· ····················· ···················· ···················· ····················
X 5322 hCoV-19/Netherlands/ZH-EMC-1048/2020|EPI_ISL_72267 ···················· ···················· ····················· ···················· ···················· ····················
X 5323 hCoV-19/Netherlands/ZH-EMC-996/2020|EPI_ISL_722674 ···················· ···················· ····················· ···················· ···················· ····················
X 5324 hCoV-19/Netherlands/GE-EMC-402/2020|EPI_ISL_722675 ···················· ···················· ····················· ···················· ···················· ····················
X 5325 hCoV-19/Netherlands/ZH-EMC-1001/2020|EPI_ISL_72267 ···················· ···················· ····················· ···················· ······T············· ····················
X 5326 hCoV-19/Netherlands/ZH-EMC-1004/2020|EPI_ISL_72267 ···················· ···················· ····················· ···················· ···················· ····················
X 5327 hCoV-19/Netherlands/ZH-EMC-1013/2020|EPI_ISL_72267 ···················· ···················· ····················· ···················· ···················· ····················
X 5328 hCoV-19/Netherlands/NB-EMC-414/2020|EPI_ISL_722679 ···················· ···················· ····················· ···················· ···················· ····················
X 5329 hCoV-19/Netherlands/un-EMC-799/2020|EPI_ISL_722680 ···················· ···················· ····················· ···················· ···················· ····················
X 5330 hCoV-19/Netherlands/UT-EMC-171/2020|EPI_ISL_722681 ···················· ···················· ····················· ···················· ···················· ····················
X 5331 hCoV-19/Netherlands/NB-EMC-442/2020|EPI_ISL_722682 ···················· ···················· ····················· ···················· ···················· ····················
X 5332 hCoV-19/Netherlands/ZH-EMC-1015/2020|EPI_ISL_72268 ···················· ···················· ····················· ···················· ······T············· ····················
X 5333 hCoV-19/Netherlands/ZH-EMC-1000/2020|EPI_ISL_72268 ···················· ···················· ····················· ···················· ······T············· ····················
X 5334 hCoV-19/Netherlands/ZH-EMC-1014/2020|EPI_ISL_72268 ···················· ···················· ····················· ···················· ······T············· ····················
X 5335 hCoV-19/Netherlands/ZH-EMC-1012/2020|EPI_ISL_72268 ···················· ···················· ····················· ···················· ······T············· ····················
X 5336 hCoV-19/Netherlands/NB-EMC-444/2020|EPI_ISL_722687 ···················· ···················· ····················· ···················· ···················· ····················
X 5337 hCoV-19/Netherlands/ZH-EMC-1029/2020|EPI_ISL_72268 ···················· ···················· ····················· ···················· ···················· ····················
X 5338 hCoV-19/Netherlands/NB-EMC-446/2020|EPI_ISL_722689 ···················· ···················· ····················· ···················· ···················· ····················
X 5339 hCoV-19/England/CAMB-1BCE53/2020|EPI_ISL_723903|20 ···················· ···················· ····················· ···················· ···················· ····················
X 5340 hCoV-19/England/CAMB-1BCE62/2020|EPI_ISL_723904|20 ···················· ···················· ····················· ···················· ···················· ····················
X 5341 hCoV-19/England/CAMB-A97936/2020|EPI_ISL_723937|20 ···················· ···················· ····················· ···················· ···················· ····················
X 5342 hCoV-19/England/CAMB-A97972/2020|EPI_ISL_723939|20 ···················· ···················· ····················· ···················· ···················· ····················
X 5343 hCoV-19/England/CAMB-A979AF/2020|EPI_ISL_723941|20 ···················· ···················· ····················· ···················· ···················· ····················
X 5344 hCoV-19/England/CAMB-A979DC/2020|EPI_ISL_723944|20 ···················· ···················· ····················· ···················· ···················· ····················
X 5345 hCoV-19/England/CAMB-A97A06/2020|EPI_ISL_723945|20 ···················· ···················· ····················· ···················· ···················· ····················
X 5346 hCoV-19/England/CAMC-B7AC3F/2020|EPI_ISL_723950|20 ···················· ···················· ····················· ···················· ···················· ····················
X 5347 hCoV-19/England/CAMC-B7AC4E/2020|EPI_ISL_723951|20 ···················· ·A·················· ····················· ···················· ···················· ··A·················
X 5348 hCoV-19/England/CAMC-B7AC5D/2020|EPI_ISL_723952|20 ···················· ···················· ····················· ···················· ···················· ····················
X 5349 hCoV-19/England/CAMC-B7AC6C/2020|EPI_ISL_723953|20 ···················· ···················· ····················· ···················· ···················· ····················
X 5350 hCoV-19/England/CAMC-B7AC7B/2020|EPI_ISL_723954|20 ···················· ···················· ····················· ···················· ···················· ····················
X 5351 hCoV-19/England/CAMC-B7ACA8/2020|EPI_ISL_723956|20 ···················· ···················· ····················· ···················· ···················· ····················
X 5352 hCoV-19/England/CAMC-B7ACE4/2020|EPI_ISL_723958|20 ···················· ···················· ····················· ···················· ···················· ····················
X 5353 hCoV-19/England/CAMC-B7AD2D/2020|EPI_ISL_723959|20 ···················· ···················· ····················· ···················· ···················· ····················
X 5354 hCoV-19/England/CAMC-B7AD5A/2020|EPI_ISL_723960|20 ···················· ···················· ····················· ···················· ···················· ····················
X 5355 hCoV-19/England/CAMC-B7ADA5/2020|EPI_ISL_723961|20 ···················· ···················· ····················· ···················· ···················· ····················
X 5356 hCoV-19/England/CAMC-B7AEA2/2020|EPI_ISL_723965|20 ···················· ···················· ····················· ···················· ···················· ····················
X 5357 hCoV-19/England/CAMC-B7AEC0/2020|EPI_ISL_723966|20 ···················· ···················· ····················· ···················· ···················· ····················
X 5358 hCoV-19/England/CAMC-B7AEDF/2020|EPI_ISL_723967|20 ···················· ···················· ····················· ···················· ···················· ····················
X 5359 hCoV-19/England/CAMC-B7AEEE/2020|EPI_ISL_723968|20 ···················· ···················· ····················· ···················· ···················· ····················
X 5360 hCoV-19/England/CAMC-B7AF18/2020|EPI_ISL_723969|20 ···················· ···················· ····················· ···················· ···················· ····················
X 5361 hCoV-19/England/CAMC-B7AF54/2020|EPI_ISL_723970|20 ···················· ···················· ····················· ···················· ···················· ····················
X 5362 hCoV-19/England/CAMC-B7AF63/2020|EPI_ISL_723971|20 ···················· ···················· ····················· ···················· ···················· ····················
X 5363 hCoV-19/England/CAMC-B7AF72/2020|EPI_ISL_723972|20 ···················· ·A·················· ····················· ···················· ···················· ····················
X 5364 hCoV-19/England/CAMC-B7AF81/2020|EPI_ISL_723973|20 ···················· ···················· ····················· ···················· ···················· ····················
X 5365 hCoV-19/England/CAMC-B7AFFA/2020|EPI_ISL_723975|20 ···················· ···················· ····················· ···················· ···················· ····················
X 5366 hCoV-19/England/CAMC-B7B005/2020|EPI_ISL_723976|20 ···················· ···················· ····················· ···················· ···················· ····················
X 5367 hCoV-19/England/CAMC-B7B014/2020|EPI_ISL_723977|20 ···················· ···················· ····················· ···················· ···················· ····················
X 5368 hCoV-19/England/CAMC-B7B041/2020|EPI_ISL_723980|20 ···················· ···················· ····················· ···················· ···················· ····················
X 5369 hCoV-19/England/CAMC-B7B050/2020|EPI_ISL_723981|20 ···················· ···················· ····················· ···················· ···················· ····················
X 5370 hCoV-19/England/CAMC-B7B102/2020|EPI_ISL_723982|20 ···················· ···················· ····················· ···················· ···················· ····················
X 5371 hCoV-19/England/CAMC-B7B111/2020|EPI_ISL_723983|20 ···················· ···················· ····················· ···················· ···················· ····················
X 5372 hCoV-19/England/CAMC-B7B120/2020|EPI_ISL_723984|20 ···················· ···················· ····················· ···················· ···················· ····················
X 5373 hCoV-19/England/CAMC-B7B14E/2020|EPI_ISL_723985|20 ···················· ···················· ····················· ···················· ···················· ····················
X 5374 hCoV-19/England/CAMC-B7B15D/2020|EPI_ISL_723986|20 ···················· ···················· ····················· ···················· ···················· ····················
X 5375 hCoV-19/England/CAMC-B7B16C/2020|EPI_ISL_723987|20 ···················· ···················· ····················· ···················· ···················· ····················
X 5376 hCoV-19/England/CAMC-B7B17B/2020|EPI_ISL_723988|20 ···················· ···················· ····················· ···················· ···················· ····················
X 5377 hCoV-19/Scotland/QEUH-C14DF7/2020|EPI_ISL_710670|2 ···················· ···················· ····················· ···················· ···················· ····················
X 5378 hCoV-19/Wales/ALDP-C06341/2020|EPI_ISL_710671|2020 ···················· ···················· ····················· ···················· ···················· ····················
X 5379 hCoV-19/Scotland/QEUH-C14911/2020|EPI_ISL_710672|2 ···················· ···················· ····················· ···················· ···················· ····················
X 5380 hCoV-19/Scotland/QEUH-C1487E/2020|EPI_ISL_710673|2 ···················· ···················· ····················· ···················· ···················· ····················

| | | | | | | | |
|---|---|---|---|---|---|---|---|
| X | 5381 hCoV-19/Scotland/QEUH-C14823/2020\|EPI_ISL_710674\|2 | .................... | .................... | .................... | .................... | .................... | .................... |
| X | 5382 hCoV-19/Scotland/QEUH-C14C54/2020\|EPI_ISL_710679\|2 | .................... | .................... | .................... | .................... | .................... | .................... |
| X | 5383 hCoV-19/Scotland/QEUH-C1695B/2020\|EPI_ISL_710680\|2 | .................... | .................... | .................... | .................... | .................... | .................... |
| X | 5384 hCoV-19/Scotland/QEUH-C168F4/2020\|EPI_ISL_710683\|2 | .................... | .................... | .................... | .................... | .................... | .................... |
| X | 5385 hCoV-19/Scotland/QEUH-C14C81/2020\|EPI_ISL_710684\|2 | .................... | .................... | .................... | .................... | .................... | .................... |
| X | 5386 hCoV-19/Scotland/QEUH-C148E7/2020\|EPI_ISL_710685\|2 | .................... | .................... | .................... | .................... | .................... | .................... |
| X | 5387 hCoV-19/Scotland/QEUH-C14850/2020\|EPI_ISL_710686\|2 | .................... | .................... | .................... | .................... | .................... | .................... |
| X | 5388 hCoV-19/Scotland/QEUH-C148AB/2020\|EPI_ISL_710687\|2 | .................... | .................... | .................... | .................... | .................... | .................... |
| X | 5389 hCoV-19/Scotland/QEUH-C1489C/2020\|EPI_ISL_710688\|2 | .................... | .................... | .................... | .................... | .................... | .................... |
| X | 5390 hCoV-19/Wales/ALDP-C06271/2020\|EPI_ISL_710689\|2020 | .................... | .................... | .................... | .................... | .................... | .................... |
| X | 5391 hCoV-19/Wales/ALDP-BB5685/2020\|EPI_ISL_710700\|2020 | .................... | .................... | .................... | .................... | .................... | .................... |
| X | 5392 hCoV-19/Wales/ALDP-BB5658/2020\|EPI_ISL_710701\|2020 | .................... | .................... | .................... | .................... | .................... | .................... |
| X | 5393 hCoV-19/Wales/ALDP-BBA046/2020\|EPI_ISL_710702\|2020 | .................... | .................... | .................... | .................... | .................... | .................... |
| X | 5394 hCoV-19/Wales/ALDP-BB4F30/2020\|EPI_ISL_710703\|2020 | .................... | .................... | .................... | .................... | .................... | .................... |
| X | 5395 hCoV-19/Scotland/ALDP-BB50A5/2020\|EPI_ISL_710704\|2 | .................... | .................... | .................... | .................... | .................... | .................... |
| X | 5396 hCoV-19/Wales/ALDP-BB4F12/2020\|EPI_ISL_710706\|2020 | .................... | .................... | .................... | .................... | .................... | .................... |
| X | 5397 hCoV-19/Wales/ALDP-BB54E5/2020\|EPI_ISL_710707\|2020 | .................... | .................... | .................... | .................... | .................... | .................... |
| X | 5398 hCoV-19/Wales/ALDP-BB552E/2020\|EPI_ISL_710708\|2020 | .................... | .................... | .................... | .................... | .................... | .................... |
| X | 5399 hCoV-19/Wales/ALDP-BB5272/2020\|EPI_ISL_710709\|2020 | .................... | .................... | .................... | .................... | .................... | .................... |
| X | 5400 hCoV-19/Wales/ALDP-BBA34D/2020\|EPI_ISL_710714\|2020 | .................... | .................... | .................... | .................... | .................... | .................... |
| X | 5401 hCoV-19/Wales/ALDP-BBA6F9/2020\|EPI_ISL_710715\|2020 | .................... | .................... | .................... | .................... | .................... | .................... |
| X | 5402 hCoV-19/Wales/ALDP-BB4EF7/2020\|EPI_ISL_710716\|2020 | .................... | .................... | .................... | .................... | .................... | .................... |
| X | 5403 hCoV-19/Wales/ALDP-BB4F4F/2020\|EPI_ISL_710717\|2020 | .................... | .................... | .................... | .................... | ....T............... | .................... |
| X | 5404 hCoV-19/Wales/ALDP-BBA35C/2020\|EPI_ISL_710718\|2020 | .................... | .................... | .................... | .................... | .................... | .................... |
| X | 5405 hCoV-19/Wales/ALDP-BBA705/2020\|EPI_ISL_710719\|2020 | .................... | .................... | .................... | .................... | .................... | .................... |
| X | 5406 hCoV-19/Wales/ALDP-BB4F03/2020\|EPI_ISL_710720\|2020 | .................... | .................... | .................... | .................... | .................... | .................... |
| X | 5407 hCoV-19/Wales/ALDP-BB4EE8/2020\|EPI_ISL_710721\|2020 | .................... | .................... | .................... | .................... | .................... | .................... |
| X | 5408 hCoV-19/Wales/ALDP-BB4A02/2020\|EPI_ISL_710722\|2020 | .................... | .................... | .................... | .................... | .................... | .................... |
| X | 5409 hCoV-19/Wales/ALDP-BB4F5E/2020\|EPI_ISL_710723\|2020 | .................... | .................... | .................... | .................... | .................... | .................... |
| X | 5410 hCoV-19/Wales/ALDP-BBA626/2020\|EPI_ISL_710724\|2020 | .................... | .................... | .................... | .................... | .................... | .................... |
| X | 5411 hCoV-19/Wales/ALDP-BBA310/2020\|EPI_ISL_710725\|2020 | .................... | .................... | .................... | .................... | .................... | .................... |
| X | 5412 hCoV-19/Wales/ALDP-BBA617/2020\|EPI_ISL_710726\|2020 | .................... | .................... | .................... | .................... | .................... | .................... |
| X | 5413 hCoV-19/Wales/ALDP-BBA714/2020\|EPI_ISL_710728\|2020 | .................... | .................... | .................... | .................... | .................... | .................... |
| X | 5414 hCoV-19/Wales/ALDP-BBA6CC/2020\|EPI_ISL_710729\|2020 | .................... | .................... | .................... | .................... | .................... | .................... |
| X | 5415 hCoV-19/Wales/ALDP-BBA2F5/2020\|EPI_ISL_710730\|2020 | .................... | .................... | .................... | .................... | .................... | .................... |
| X | 5416 hCoV-19/Wales/ALDP-BB4ED9/2020\|EPI_ISL_710731\|2020 | .................... | .................... | .................... | .................... | .................... | .................... |
| X | 5417 hCoV-19/Wales/ALDP-BB553D/2020\|EPI_ISL_710732\|2020 | .................... | .................... | .................... | .................... | .................... | .................... |
| X | 5418 hCoV-19/Wales/ALDP-BBA83F/2020\|EPI_ISL_710733\|2020 | .................... | .................... | .................... | .................... | .................... | .................... |
| X | 5419 hCoV-19/Wales/ALDP-BB94D2/2020\|EPI_ISL_710734\|2020 | .................... | .................... | .................... | .................... | .................... | .................... |
| X | 5420 hCoV-19/Wales/ALDP-BB9830/2020\|EPI_ISL_710735\|2020 | .................... | .................... | .................... | .................... | .................... | .................... |
| X | 5421 hCoV-19/Wales/ALDP-BB9DB9/2020\|EPI_ISL_710736\|2020 | .................... | .................... | .................... | .................... | .................... | .................... |
| X | 5422 hCoV-19/Wales/ALDP-BB549A/2020\|EPI_ISL_710737\|2020 | .................... | .................... | .................... | .................... | .................... | .................... |
| X | 5423 hCoV-19/Wales/ALDP-BB985E/2020\|EPI_ISL_710738\|2020 | .................... | .................... | .................... | .................... | .................... | .................... |
| X | 5424 hCoV-19/Wales/ALDP-BB9D22/2020\|EPI_ISL_710739\|2020 | .................... | .................... | .................... | .................... | .................... | .................... |
| X | 5425 hCoV-19/Wales/ALDP-BB940F/2020\|EPI_ISL_710740\|2020 | .................... | .................... | .................... | .................... | .................... | .................... |
| X | 5426 hCoV-19/Wales/ALDP-BBA8D5/2020\|EPI_ISL_710741\|2020 | .................... | .................... | .................... | .................... | .................... | ...A................ |
| X | 5427 hCoV-19/Wales/ALDP-BB943C/2020\|EPI_ISL_710742\|2020 | .................... | .................... | .................... | .................... | .................... | .................... |
| X | 5428 hCoV-19/Wales/ALDP-BB9DC8/2020\|EPI_ISL_710744\|2020 | .................... | .................... | .................... | .................... | .................... | .................... |
| X | 5429 hCoV-19/Scotland/QEUH-BCC2A5/2020\|EPI_ISL_710768\|2 | .................... | .................... | .................... | .................... | .................... | .................... |
| X | 5430 hCoV-19/Scotland/QEUH-BC38E8/2020\|EPI_ISL_710772\|2 | .................... | .................... | .................... | .................... | .................... | .................... |
| X | 5431 hCoV-19/Scotland/QEUH-BCC3A2/2020\|EPI_ISL_710774\|2 | .................... | .................... | .................... | .................... | .................... | .................... |
| X | 5432 hCoV-19/Scotland/QEUH-BCC2F0/2020\|EPI_ISL_710775\|2 | .................... | .................... | .................... | .................... | .................... | .................... |
| X | 5433 hCoV-19/Scotland/QEUH-BCC25A/2020\|EPI_ISL_710776\|2 | .................... | .................... | .................... | .................... | .................... | .................... |
| X | 5434 hCoV-19/Scotland/QEUH-BC395E/2020\|EPI_ISL_710777\|2 | .................... | .................... | .................... | .................... | .................... | .................... |
| X | 5435 hCoV-19/Denmark/DCGC-15932/2020\|EPI_ISL_714259\|202 | ........T........... | .................... | .................... | .................... | .................... | .................... |
| X | 5436 hCoV-19/Denmark/DCGC-15961/2020\|EPI_ISL_714260\|202 | .................... | .................... | .................... | .................... | .................... | .................... |
| X | 5437 hCoV-19/Denmark/DCGC-16101/2020\|EPI_ISL_714261\|202 | .................... | .................... | .................... | .................... | .................... | .................... |
| X | 5438 hCoV-19/Denmark/DCGC-16127/2020\|EPI_ISL_714262\|202 | .................... | .................... | .................... | .................... | .................... | .................... |
| X | 5439 hCoV-19/Denmark/DCGC-16159/2020\|EPI_ISL_714263\|202 | .................... | .................... | .................... | .................... | .................... | .................... |
| X | 5440 hCoV-19/Denmark/DCGC-15931/2020\|EPI_ISL_714264\|202 | .................... | .................... | .................... | .................... | .................... | .................... |
| X | 5441 hCoV-19/Denmark/DCGC-16064/2020\|EPI_ISL_714265\|202 | .................... | .................... | .................... | .................... | .................... | .................... |
| X | 5442 hCoV-19/Denmark/DCGC-16080/2020\|EPI_ISL_714266\|202 | .................... | .................... | .................... | .................... | .................... | .................... |
| X | 5443 hCoV-19/Denmark/DCGC-16129/2020\|EPI_ISL_714267\|202 | .................... | .................... | .................... | .................... | .................... | .................... |
| X | 5444 hCoV-19/Denmark/DCGC-16317/2020\|EPI_ISL_714268\|202 | .................... | .................... | .................... | .................... | .................... | .................... |
| X | 5445 hCoV-19/Denmark/DCGC-16695/2020\|EPI_ISL_714269\|202 | .................... | .................... | .................... | .................... | .................... | .................... |
| X | 5446 hCoV-19/Denmark/DCGC-16604/2020\|EPI_ISL_714270\|202 | .................... | .A.................. | .................... | .................... | .................... | ...A................ |
| X | 5447 hCoV-19/Denmark/DCGC-16309/2020\|EPI_ISL_714271\|202 | .................... | .................... | .................... | .................... | .................... | .................... |
| X | 5448 hCoV-19/Denmark/DCGC-16028/2020\|EPI_ISL_714289\|202 | .................... | .................... | .................... | .................... | .................... | .................... |
| X | 5449 hCoV-19/Denmark/DCGC-16108/2020\|EPI_ISL_714290\|202 | .................... | .................... | .................... | .................... | .................... | .................... |
| X | 5450 hCoV-19/Denmark/DCGC-16167/2020\|EPI_ISL_714291\|202 | .................... | .................... | .................... | .................... | .................... | .................... |
| X | 5451 hCoV-19/Denmark/DCGC-16269/2020\|EPI_ISL_714292\|202 | .................... | .................... | .................... | .................... | .................... | .................... |
| X | 5452 hCoV-19/Denmark/DCGC-16272/2020\|EPI_ISL_714293\|202 | .................... | .A.................. | .................... | .................... | .................... | T................... |
| X | 5453 hCoV-19/Denmark/DCGC-16307/2020\|EPI_ISL_714294\|202 | .................... | .................... | .................... | .................... | .................... | .................... |
| X | 5454 hCoV-19/Denmark/DCGC-16356/2020\|EPI_ISL_714295\|202 | .................... | .................... | .................... | .................... | .................... | .................... |
| X | 5455 hCoV-19/Denmark/DCGC-16427/2020\|EPI_ISL_714296\|202 | .................... | .................... | .................... | .................... | .................... | .................... |
| X | 5456 hCoV-19/Denmark/DCGC-16464/2020\|EPI_ISL_714297\|202 | .................... | .................... | .................... | .................... | .................... | .................... |
| X | 5457 hCoV-19/Denmark/DCGC-16665/2020\|EPI_ISL_714298\|202 | .................... | .................... | .................... | .................... | .................... | .................... |
| X | 5458 hCoV-19/England/CAMC-C43C6B/2020\|EPI_ISL_719011\|20 | .................... | .................... | .................... | .................... | .................... | .................... |
| X | 5459 hCoV-19/England/CAMC-C3EFD3/2020\|EPI_ISL_719012\|20 | .................... | .................... | .................... | .................... | .................... | .................... |
| X | 5460 hCoV-19/England/CAMC-C40E86/2020\|EPI_ISL_719019\|20 | .................... | .................... | .................... | .................... | .................... | .................... |

| | | | | | | | | |
|---|---|---|---|---|---|---|---|---|
| X | 5461 | hCoV-19/England/CAMC-C3EFC4/2020\|EPI_ISL_719040\|20 | ···················· | ···················· | ····················· | ···················· | ···················· | ···················· |
| X | 5462 | hCoV-19/England/CAMC-C44EEC/2020\|EPI_ISL_719044\|20 | ···················· | ···················· | ····················· | ···················· | ···················· | ···················· |
| X | 5463 | hCoV-19/England/CAMC-C42B32/2020\|EPI_ISL_719048\|20 | ···················· | ·A·················· | ····················· | ···················· | ···················· | ···A················ |
| X | 5464 | hCoV-19/England/CAMC-C42B41/2020\|EPI_ISL_719053\|20 | ············T······· | ···················· | ····················· | ···················· | ···················· | ···················· |
| X | 5465 | hCoV-19/England/CAMC-C436D6/2020\|EPI_ISL_719054\|20 | ···········T········ | ···················· | ····················· | ···················· | ···················· | ···················· |
| X | 5466 | hCoV-19/England/CAMC-C3FAA4/2020\|EPI_ISL_719078\|20 | ···················· | ···················· | ····················· | ···················· | ···················· | ···················· |
| X | 5467 | hCoV-19/England/CAMC-C43AAD/2020\|EPI_ISL_719086\|20 | ···················· | ···················· | ····················· | ···················· | ···················· | ···················· |
| X | 5468 | hCoV-19/England/CAMC-C40BF8/2020\|EPI_ISL_719090\|20 | ···················· | ···················· | ····················· | ···················· | ······T············· | ···················· |
| X | 5469 | hCoV-19/England/CAMC-C40E2C/2020\|EPI_ISL_719091\|20 | ···················· | ···················· | ····················· | ···················· | ···················· | ···················· |
| X | 5470 | hCoV-19/England/CAMC-C41C5E/2020\|EPI_ISL_719096\|20 | ···················· | ···················· | ····················· | ···················· | ···················· | ···················· |
| X | 5471 | hCoV-19/England/CAMC-C40E68/2020\|EPI_ISL_719099\|20 | ···················· | ···················· | ····················· | ···················· | ······T············· | ···················· |
| X | 5472 | hCoV-19/England/CAMC-C3FA59/2020\|EPI_ISL_719103\|20 | ···················· | ···················· | ····················· | ···················· | ······T············· | ···················· |
| X | 5473 | hCoV-19/England/CAMC-C3F057/2020\|EPI_ISL_719104\|20 | ···················· | ···················· | ····················· | ···················· | ······T············· | ···················· |
| X | 5474 | hCoV-19/England/CAMC-C412E3/2020\|EPI_ISL_719107\|20 | ···················· | ···················· | ····················· | ···················· | ···················· | ···················· |
| X | 5475 | hCoV-19/England/MILK-BB247F/2020\|EPI_ISL_720002\|20 | ···················· | ···················· | ····················· | ···················· | ···················· | ···················· |
| X | 5476 | hCoV-19/England/MILK-BB08EE/2020\|EPI_ISL_720006\|20 | ···················· | ···················· | ····················· | ···················· | ···················· | ···················· |
| X | 5477 | hCoV-19/England/MILK-BB02E0/2020\|EPI_ISL_720030\|20 | ···················· | ···················· | ····················· | ···················· | ···················· | ···················· |
| X | 5478 | hCoV-19/England/MILK-BB024A/2020\|EPI_ISL_720033\|20 | ···················· | ···················· | ····················· | ···················· | ···················· | ···················· |
| X | 5479 | hCoV-19/England/MILK-BB23FA/2020\|EPI_ISL_720034\|20 | ···················· | ···················· | ····················· | ···················· | ···················· | ···················· |
| X | 5480 | hCoV-19/England/MILK-BB1795/2020\|EPI_ISL_720036\|20 | ···················· | ·A·················· | ····················· | ···················· | ···················· | ···A················ |
| X | 5481 | hCoV-19/England/MILK-BB1E81/2020\|EPI_ISL_720038\|20 | ···················· | ···················· | ····················· | ···················· | ···················· | ···················· |
| X | 5482 | hCoV-19/England/MILK-BB08DF/2020\|EPI_ISL_720039\|20 | ···················· | ···················· | ····················· | ···················· | ···················· | ···················· |
| X | 5483 | hCoV-19/England/MILK-BB20F3/2020\|EPI_ISL_720042\|20 | ···················· | ···················· | ····················· | ···················· | ······T············· | ···················· |
| X | 5484 | hCoV-19/England/MILK-BB2099/2020\|EPI_ISL_720043\|20 | ···················· | ···················· | ····················· | ···················· | ···················· | ···················· |
| X | 5485 | hCoV-19/England/MILK-BB1E54/2020\|EPI_ISL_720044\|20 | ···················· | ···················· | ····················· | ···················· | ···················· | ···················· |
| X | 5486 | hCoV-19/England/MILK-BB0268/2020\|EPI_ISL_720046\|20 | ···················· | ···················· | ····················· | ···················· | ···················· | ···A················ |
| X | 5487 | hCoV-19/England/MILK-BB1892/2020\|EPI_ISL_720047\|20 | ···················· | ···················· | ····················· | ···················· | ···················· | ···················· |
| X | 5488 | hCoV-19/England/MILK-BB1F8E/2020\|EPI_ISL_720048\|20 | ···················· | ···················· | ····················· | ···················· | ···················· | ···················· |
| X | 5489 | hCoV-19/England/MILK-BB157D/2020\|EPI_ISL_720049\|20 | ···················· | ···················· | ····················· | ···················· | ···················· | ···················· |
| X | 5490 | hCoV-19/England/MILK-BB02FF/2020\|EPI_ISL_720050\|20 | ···················· | ···················· | ····················· | ···················· | ···················· | ···················· |
| X | 5491 | hCoV-19/England/MILK-BB25B8/2020\|EPI_ISL_720053\|20 | ···················· | ···················· | ····················· | ···················· | ···················· | ···················· |
| X | 5492 | hCoV-19/England/MILK-BB16C5/2020\|EPI_ISL_720054\|20 | ···················· | ···················· | ····················· | ···················· | ···················· | ···················· |
| X | 5493 | hCoV-19/England/MILK-BB122B/2020\|EPI_ISL_720056\|20 | ···················· | ···················· | ····················· | ···················· | ···················· | ···················· |
| X | 5494 | hCoV-19/England/MILK-BB263D/2020\|EPI_ISL_720058\|20 | ···················· | ···················· | ····················· | ···················· | ···················· | ···················· |
| X | 5495 | hCoV-19/England/MILK-BB0277/2020\|EPI_ISL_720060\|20 | ···················· | ···················· | ····················· | ···················· | ···················· | ···················· |
| X | 5496 | hCoV-19/England/MILK-BB1355/2020\|EPI_ISL_720063\|20 | ···················· | ···················· | ····················· | ···················· | ···················· | ···················· |
| X | 5497 | hCoV-19/England/MILK-BB1777/2020\|EPI_ISL_720064\|20 | ···················· | ···················· | ····················· | ···················· | ···················· | ···················· |
| X | 5498 | hCoV-19/England/MILK-BB1382/2020\|EPI_ISL_720067\|20 | ···················· | ···················· | ····················· | ···················· | ···················· | ···················· |
| X | 5499 | hCoV-19/England/MILK-BB18CF/2020\|EPI_ISL_720069\|20 | ···················· | ···················· | ····················· | ···················· | ···················· | ···················· |
| X | 5500 | hCoV-19/England/MILK-BB1452/2020\|EPI_ISL_720070\|20 | ···················· | ···················· | ····················· | ···················· | ···················· | ···················· |
| X | 5501 | hCoV-19/England/MILK-BB155F/2020\|EPI_ISL_720072\|20 | ···················· | ···················· | ····················· | ···················· | ···················· | ···················· |
| X | 5502 | hCoV-19/England/MILK-BB148F/2020\|EPI_ISL_720073\|20 | ···················· | ···················· | ····················· | ···················· | ···················· | ···················· |
| X | 5503 | hCoV-19/England/MILK-BB0505/2020\|EPI_ISL_720074\|20 | ···················· | ···················· | ····················· | ···················· | ···················· | ···················· |
| X | 5504 | hCoV-19/England/MILK-BB13EC/2020\|EPI_ISL_720075\|20 | ···················· | ···················· | ····················· | ···················· | ···A················ | ···················· |
| X | 5505 | hCoV-19/England/MILK-BB17D1/2020\|EPI_ISL_720077\|20 | ···················· | ···················· | ····················· | ···················· | ···················· | ···················· |
| X | 5506 | hCoV-19/England/MILK-BB1285/2020\|EPI_ISL_720078\|20 | ···················· | ···················· | ····················· | ···················· | ···················· | ···················· |
| X | 5507 | hCoV-19/England/MILK-BB13DD/2020\|EPI_ISL_720079\|20 | ···················· | ···················· | ····················· | ···················· | ···················· | ···················· |
| X | 5508 | hCoV-19/England/MILK-BB149E/2020\|EPI_ISL_720084\|20 | ···················· | ···················· | ····················· | ···················· | ···················· | ···················· |
| X | 5509 | hCoV-19/England/MILK-BB25A9/2020\|EPI_ISL_720086\|20 | ···················· | ···················· | ····················· | ···················· | ···················· | ···················· |
| X | 5510 | hCoV-19/England/MILK-BB1F42/2020\|EPI_ISL_720087\|20 | ···················· | ···················· | ····················· | ···················· | ···················· | ···················· |
| X | 5511 | hCoV-19/England/MILK-BB0514/2020\|EPI_ISL_720089\|20 | ···················· | ···················· | ····················· | ···················· | ···················· | ···················· |
| X | 5512 | hCoV-19/England/MILK-BB01E3/2020\|EPI_ISL_720090\|20 | ···················· | ···················· | ····················· | ···················· | ···················· | ···················· |
| X | 5513 | hCoV-19/England/MILK-BB1BC6/2020\|EPI_ISL_720091\|20 | ···················· | ···················· | ····················· | ···················· | ···················· | ···················· |
| X | 5514 | hCoV-19/England/MILK-BB0550/2020\|EPI_ISL_720093\|20 | ···················· | ···················· | ····················· | ···················· | ···················· | ···················· |
| X | 5515 | hCoV-19/England/MILK-BB020E/2020\|EPI_ISL_720094\|20 | ···················· | ···················· | ····················· | ···················· | ···················· | ···················· |
| X | 5516 | hCoV-19/England/MILK-BB0981/2020\|EPI_ISL_720098\|20 | ···················· | ···················· | ····················· | ···················· | ···················· | ···················· |
| X | 5517 | hCoV-19/England/MILK-BB1C2D/2020\|EPI_ISL_720099\|20 | ···················· | ···················· | ····················· | ···················· | ···················· | ···················· |
| X | 5518 | hCoV-19/England/ALDP-BC8591/2020\|EPI_ISL_721316\|20 | ···················· | ···················· | ····················· | ···················· | ···················· | ···················· |
| X | 5519 | hCoV-19/England/ALDP-BC878C/2020\|EPI_ISL_721317\|20 | ···················· | ···················· | ····················· | ···················· | ···················· | ···················· |
| X | 5520 | hCoV-19/England/ALDP-BC87B9/2020\|EPI_ISL_721318\|20 | ···················· | ···················· | ····················· | ···················· | ···················· | ···················· |
| X | 5521 | hCoV-19/England/ALDP-BC885C/2020\|EPI_ISL_721319\|20 | ···················· | ···················· | ····················· | ···················· | ···················· | ···················· |
| X | 5522 | hCoV-19/England/ALDP-BC7D5E/2020\|EPI_ISL_721327\|20 | ···················· | ···················· | ····················· | ···················· | ···················· | ···················· |
| X | 5523 | hCoV-19/England/ALDP-BC7B72/2020\|EPI_ISL_721328\|20 | ···················· | ···················· | ····················· | ···················· | ···················· | ···················· |
| X | 5524 | hCoV-19/England/ALDP-BC86AD/2020\|EPI_ISL_721329\|20 | ···················· | ···················· | ····················· | ···················· | ···················· | ···················· |
| X | 5525 | hCoV-19/England/ALDP-BC6548/2020\|EPI_ISL_721331\|20 | ···················· | ···················· | ····················· | ···················· | ···················· | ···················· |
| X | 5526 | hCoV-19/England/ALDP-BC633F/2020\|EPI_ISL_721332\|20 | ···················· | ···················· | ····················· | ···················· | ···················· | ···················· |
| X | 5527 | hCoV-19/England/ALDP-BC635D/2020\|EPI_ISL_721333\|20 | ···················· | ···················· | ····················· | ···················· | ···················· | ···················· |
| X | 5528 | hCoV-19/England/ALDP-BC7583/2020\|EPI_ISL_721334\|20 | ···················· | ···················· | ····················· | ···················· | ···················· | ···················· |
| X | 5529 | hCoV-19/England/ALDP-BC88D4/2020\|EPI_ISL_721336\|20 | ···················· | ···················· | ····················· | ···················· | ···················· | ···················· |
| X | 5530 | hCoV-19/England/ALDP-BC73C5/2020\|EPI_ISL_721337\|20 | ···················· | ···················· | ····················· | ···················· | ···················· | ···················· |
| X | 5531 | hCoV-19/England/ALDP-BC84FE/2020\|EPI_ISL_721360\|20 | ···················· | ···················· | ····················· | ···················· | ···················· | ···················· |
| X | 5532 | hCoV-19/England/ALDP-BC6241/2020\|EPI_ISL_721361\|20 | ···················· | ···················· | ····················· | ···················· | ···················· | ···················· |
| X | 5533 | hCoV-19/England/ALDP-BC79D2/2020\|EPI_ISL_721364\|20 | ···················· | ···················· | ····················· | ···················· | ···················· | ···················· |
| X | 5534 | hCoV-19/England/ALDP-BC6320/2020\|EPI_ISL_721365\|20 | ···················· | ···················· | ····················· | ···················· | ······T············· | ···················· |
| X | 5535 | hCoV-19/England/ALDP-BC62E7/2020\|EPI_ISL_721366\|20 | ···················· | ···················· | ····················· | ···················· | ···················· | ···················· |
| X | 5536 | hCoV-19/England/ALDP-BC65DF/2020\|EPI_ISL_721367\|20 | ···················· | ···················· | ····················· | ···················· | ···················· | ···················· |
| X | 5537 | hCoV-19/England/ALDP-BC7C60/2020\|EPI_ISL_721368\|20 | ···················· | ···················· | ····················· | ···················· | ···················· | ···················· |
| X | 5538 | hCoV-19/England/ALDP-BC7A48/2020\|EPI_ISL_721369\|20 | ···················· | ···················· | ····················· | ···················· | ···················· | ···················· |
| X | 5539 | hCoV-19/England/ALDP-BC64E1/2020\|EPI_ISL_721370\|20 | ···················· | ···················· | ····················· | ···················· | ···················· | ···················· |
| X | 5540 | hCoV-19/England/ALDP-BC7495/2020\|EPI_ISL_721371\|20 | ···················· | ···················· | ····················· | ···················· | ···················· | ···················· |

X 5541 hCoV-19/England/ALDP-BC6311/2020|EPI_ISL_721373|20 .................... .................... .................... .................... .................... ....................
X 5542 hCoV-19/England/ALDP-BC7978/2020|EPI_ISL_721374|20 .................... .................... .................... .................... .................... ....................
X 5543 hCoV-19/England/ALDP-BC61CC/2020|EPI_ISL_721375|20 .................... .................... .................... .................... .................... ....................
X 5544 hCoV-19/England/ALDP-BC6654/2020|EPI_ISL_721376|20 .................... .................... .................... .................... .................... ....................
X 5545 hCoV-19/England/ALDP-BC65FD/2020|EPI_ISL_721377|20 .................... .................... .................... .................... .................... ....................
X 5546 hCoV-19/England/ALDP-BC87D7/2020|EPI_ISL_721378|20 .................... .................... .................... .................... .................... ....................
X 5547 hCoV-19/England/ALDP-BC8801/2020|EPI_ISL_721379|20 .................... .................... .................... .................... .................... ....................
X 5548 hCoV-19/England/ALDP-BC89E0/2020|EPI_ISL_721380|20 .................... .................... .................... .................... .................... ....................
X 5549 hCoV-19/England/ALDP-BC884D/2020|EPI_ISL_721381|20 .................... .................... .................... .................... .................... ....................
X 5550 hCoV-19/England/ALDP-BC629C/2020|EPI_ISL_721382|20 .................... .................... .................... .................... .................... ....................
X 5551 hCoV-19/England/ALDP-BC67AC/2020|EPI_ISL_721383|20 .................... .................... .................... .................... .................... ....................
X 5552 hCoV-19/England/ALDP-BC7B45/2020|EPI_ISL_721384|20 .................... .................... .................... .................... .................... ....................
X 5553 hCoV-19/England/ALDP-BC7BFA/2020|EPI_ISL_721385|20 .................... .................... .................... .................... .................... ....................
X 5554 hCoV-19/England/ALDP-BC75DE/2020|EPI_ISL_721387|20 .................... .................... .................... .................... .................... ....................
X 5555 hCoV-19/England/ALDP-BC7AFD/2020|EPI_ISL_721388|20 .................... .................... .................... .................... .................... ....................
X 5556 hCoV-19/England/ALDP-BC63A8/2020|EPI_ISL_721391|20 .................... .................... .................... .................... .................... ....................
X 5557 hCoV-19/England/ALDP-BC77E7/2020|EPI_ISL_721392|20 .................... .................... .................... .................... .................... ....................
X 5558 hCoV-19/England/ALDP-BC869E/2020|EPI_ISL_721393|20 .................... .................... .................... .................... .................... ....................
X 5559 hCoV-19/England/ALDP-BC7486/2020|EPI_ISL_721394|20 .................... .................... .................... .................... .................... ....................
X 5560 hCoV-19/England/ALDP-BC7CF7/2020|EPI_ISL_721396|20 .................... .................... .................... .................... .................... ....................
X 5561 hCoV-19/England/ALDP-BC868F/2020|EPI_ISL_721397|20 .................... .................... .................... .................... .................... ....................
X 5562 hCoV-19/England/ALDP-BC8528/2020|EPI_ISL_721398|20 .................... .................... .................... .................... .................... ....................
X 5563 hCoV-19/England/ALDP-BC89C2/2020|EPI_ISL_721420|20 .................... .................... .................... .................... .................... ....................
X 5564 hCoV-19/England/ALDP-BC85BF/2020|EPI_ISL_721423|20 .................... .................... .................... .................... .................... ....................
X 5565 hCoV-19/England/ALDP-BC777E/2020|EPI_ISL_721424|20 .................... .................... .................... .................... .................... ....................
X 5566 hCoV-19/England/ALDP-BC74C2/2020|EPI_ISL_721425|20 .................... .................... .................... .................... .....T.............. ....................
X 5567 hCoV-19/Netherlands/UT-EMC-195/2020|EPI_ISL_722647 .................... .................... .................... .................... .................... ....................
X 5568 hCoV-19/Netherlands/GE-EMC-351/2020|EPI_ISL_722648 .................... .................... .................... .................... .................... ....................
X 5569 hCoV-19/Netherlands/GE-EMC-352/2020|EPI_ISL_722649 .................... .................... .................... .................... .................... ....................
X 5570 hCoV-19/Netherlands/ZH-EMC-1026/2020|EPI_ISL_72265 .................... .................... .................... .................... .................... ....................
X 5571 hCoV-19/Netherlands/ZH-EMC-1027/2020|EPI_ISL_72265 .................... .................... .................... .................... .....T.............. ....................
X 5572 hCoV-19/Netherlands/ZH-EMC-983/2020|EPI_ISL_722660 .................... .................... .................... .................... .....T.............. ....................
X 5573 hCoV-19/Netherlands/ZH-EMC-990/2020|EPI_ISL_722661 .................... .................... .................... .................... .................... ....................
X 5574 hCoV-19/Netherlands/UT-EMC-205/2020|EPI_ISL_722662 .................... .................... .................... .................... .................... ....................
X 5575 hCoV-19/Netherlands/ZH-EMC-970/2020|EPI_ISL_722663 .................... .................... .................... .................... .................... ....................
X 5576 hCoV-19/Netherlands/ZH-EMC-973/2020|EPI_ISL_722664 .................... .................... .................... .................... .....T.............. ....................
X 5577 hCoV-19/Netherlands/ZH-EMC-975/2020|EPI_ISL_722665 .................... .................... .................... .................... .....T.............. ....................
X 5578 hCoV-19/Netherlands/ZH-EMC-977/2020|EPI_ISL_722666 .................... .................... .................... .................... .....T.............. ....................
X 5579 hCoV-19/Netherlands/GE-EMC-407/2020|EPI_ISL_722667 .................... .................... .................... .................... .................... ....................
X 5580 hCoV-19/Netherlands/NB-EMC-440/2020|EPI_ISL_722668 .................... .................... .................... .................... .................... ....................
X 5581 hCoV-19/Netherlands/UT-EMC-217/2020|EPI_ISL_722690 .................... .................... .................... .................... .................... ....................
X 5582 hCoV-19/Netherlands/ZH-EMC-1036/2020|EPI_ISL_72269 .................... .................... .................... .................... .....T.............. ....................
X 5583 hCoV-19/Netherlands/ZH-EMC-1043/2020|EPI_ISL_72269 .................... .................... .................... .................... .................... ....................
X 5584 hCoV-19/Netherlands/GE-EMC-421/2020|EPI_ISL_722693 .................... .................... .................... .................... .................... ....................
X 5585 hCoV-19/Netherlands/NB-EMC-447/2020|EPI_ISL_722694 .................... .................... .................... .................... .................... ....................
X 5586 hCoV-19/Netherlands/UT-EMC-218/2020|EPI_ISL_722695 .................... .................... .................... .................... .................... ....................
X 5587 hCoV-19/Netherlands/ZH-EMC-1044/2020|EPI_ISL_72269 .................... .................... .................... .................... .................... ....................
X 5588 hCoV-19/Netherlands/ZH-EMC-1045/2020|EPI_ISL_72269 .................... .................... .................... .................... .................... ....................
X 5589 hCoV-19/Netherlands/LI-EMC-116/2020|EPI_ISL_722698 .................... .................... .................... .................... .................... ...............A....
X 5590 hCoV-19/Netherlands/NB-EMC-362/2020|EPI_ISL_722699 .................... .................... .................... .................... .................... ....................
X 5591 hCoV-19/Netherlands/NB-EMC-364/2020|EPI_ISL_722700 .................... .................... .................... .................... .................... ....................
X 5592 hCoV-19/Netherlands/UT-EMC-167/2020|EPI_ISL_722701 .................... .................... .................... .................... .................... ....................
X 5593 hCoV-19/Netherlands/ZH-EMC-907/2020|EPI_ISL_722702 .................... .................... .................... .................... .................... ....................
X 5594 hCoV-19/Netherlands/ZH-EMC-908/2020|EPI_ISL_722703 .................... .................... .................... .................... .................... ....................
X 5595 hCoV-19/Netherlands/ZH-EMC-909/2020|EPI_ISL_722704 .................... .................... .................... .................... .................... ....................
X 5596 hCoV-19/Netherlands/ZH-EMC-910/2020|EPI_ISL_722705 .................... .................... .................... .................... .................... ....................
X 5597 hCoV-19/Netherlands/ZH-EMC-912/2020|EPI_ISL_722706 .................... .................... .................... .................... .................... ....................
X 5598 hCoV-19/Netherlands/ZH-EMC-913/2020|EPI_ISL_722707 .................... .................... .................... .................... .....T.............. ....................
X 5599 hCoV-19/Netherlands/ZH-EMC-916/2020|EPI_ISL_722708 .................... .................... .................... .................... .................... ....................
X 5600 hCoV-19/Netherlands/ZH-EMC-967/2020|EPI_ISL_722709 .................... .................... .................... .................... .................... ....................
X 5601 hCoV-19/Netherlands/ZH-EMC-968/2020|EPI_ISL_722710 .................... .................... .................... .................... .................... ....................
X 5602 hCoV-19/Netherlands/GE-EMC-422/2020|EPI_ISL_722711 .................... .................... .................... .................... .................... ....................
X 5603 hCoV-19/Netherlands/GE-EMC-431/2020|EPI_ISL_722712 .................... .................... .................... .................... .................... ....................
X 5604 hCoV-19/Netherlands/NB-EMC-450/2020|EPI_ISL_722713 .................... .................... .................... .................... .................... ....................
X 5605 hCoV-19/Netherlands/FL-EMC-41/2020|EPI_ISL_722714| .................... .................... .................... .................... .................... ....................
X 5606 hCoV-19/Netherlands/GE-EMC-359/2020|EPI_ISL_722715 .................... .................... .................... .................... .................... ....................
X 5607 hCoV-19/Netherlands/GE-EMC-360/2020|EPI_ISL_722716 .................... .................... .................... .................... .....T.............. ....................
X 5608 hCoV-19/Netherlands/GE-EMC-361/2020|EPI_ISL_722717 .................... .................... .................... .................... .................... ....................
X 5609 hCoV-19/Netherlands/LI-EMC-114/2020|EPI_ISL_722718 .................... .................... .................... .................... .................... ....................
X 5610 hCoV-19/Netherlands/LI-EMC-115/2020|EPI_ISL_722719 .................... .................... .................... .................... .................... ....................
X 5611 hCoV-19/Netherlands/NB-EMC-363/2020|EPI_ISL_722720 .................... .................... .................... .................... .................... ....................
X 5612 hCoV-19/Netherlands/UT-EMC-201/2020|EPI_ISL_722721 .................... .................... .................... .................... .....T.............. ....................
X 5613 hCoV-19/Netherlands/ZH-EMC-914/2020|EPI_ISL_722722 .................... .................... .................... .................... .................... ....................
X 5614 hCoV-19/Netherlands/ZH-EMC-917/2020|EPI_ISL_722723 .................... .................... .................... .................... .................... ....................
X 5615 hCoV-19/Netherlands/GE-EMC-353/2020|EPI_ISL_722750 .................... .................... .................... .................... .................... ....................
X 5616 hCoV-19/Netherlands/NB-EMC-433/2020|EPI_ISL_722751 .................... .................... .................... .................... .................... ....................
X 5617 hCoV-19/Netherlands/NH-EMC-95/2020|EPI_ISL_722752| .................... .................... .................... .................... .................... ....................
X 5618 hCoV-19/Netherlands/UT-EMC-174/2020|EPI_ISL_722753 .................... .................... .................... .................... .................... ....................
X 5619 hCoV-19/Netherlands/UT-EMC-176/2020|EPI_ISL_722754 .................... .................... .................... .................... .................... ....................
X 5620 hCoV-19/Netherlands/ZH-EMC-937/2020|EPI_ISL_722755 .................... .................... .................... .................... .................... ....................

X 5621 hCoV-19/Netherlands/ZH-EMC-982/2020|EPI_ISL_722756 ···················· ···················· ···················· ···················· ······T············· ····················
X 5622 hCoV-19/England/CAMC-B7B199/2020|EPI_ISL_723989|20 ···················· ···················· ···················· ···················· ···················· ····················
X 5623 hCoV-19/England/CAMC-B7B1B7/2020|EPI_ISL_723990|20 ···················· ···················· ···················· ···················· ···················· ····················
X 5624 hCoV-19/England/CAMC-B7B1C6/2020|EPI_ISL_723991|20 ···················· ···················· ···················· ···················· ···················· ····················
X 5625 hCoV-19/England/CAMC-B7B1D5/2020|EPI_ISL_723992|20 ···················· ···················· ···················· ···················· ···················· ····················
X 5626 hCoV-19/England/CAMC-B7B1E4/2020|EPI_ISL_723993|20 ···················· ···················· ···················· ···················· ···················· ····················
X 5627 hCoV-19/England/CAMC-B7B20F/2020|EPI_ISL_723994|20 ···················· ···················· ···················· ···················· ···················· ····················
X 5628 hCoV-19/England/CAMC-B7B21E/2020|EPI_ISL_723995|20 ···················· ···················· ···················· ···················· ···················· ····················
X 5629 hCoV-19/England/CAMC-B7B22D/2020|EPI_ISL_723996|20 ···················· ···················· ···················· ···················· ···················· ····················
X 5630 hCoV-19/England/CAMC-B7B24B/2020|EPI_ISL_723997|20 ···················· ···················· ···················· ···················· ···················· ····················
X 5631 hCoV-19/England/CAMC-B7B2A5/2020|EPI_ISL_723998|20 ···················· ···················· ···················· ···················· ···················· ····················
X 5632 hCoV-19/England/CAMC-B7B2B4/2020|EPI_ISL_723999|20 ···················· ···················· ···················· ···················· ······T············· ····················
X 5633 hCoV-19/Wales/ALDP-BCAACD/2020|EPI_ISL_710745|2020 ···················· ···················· ···················· ···················· ···················· ····················
X 5634 hCoV-19/Wales/ALDP-BCAB15/2020|EPI_ISL_710747|2020 ···················· ···················· ···················· ···················· ···················· ····················
X 5635 hCoV-19/Scotland/QEUH-BC4656/2020|EPI_ISL_710748|2 ···················· ···················· ···················· ···················· ···················· ····················
X 5636 hCoV-19/Wales/ALDP-BCAAEB/2020|EPI_ISL_710749|2020 ···················· ···················· ···················· ···················· ···················· ····················
X 5637 hCoV-19/Wales/ALDP-BCA7C6/2020|EPI_ISL_710750|2020 ···················· ···················· ···················· ···················· ···················· ····················
X 5638 hCoV-19/Wales/ALDP-BCAB24/2020|EPI_ISL_710751|2020 ···················· ···················· ···················· ···················· ···················· ····················
X 5639 hCoV-19/Scotland/QEUH-BC39B8/2020|EPI_ISL_710753|2 ···················· ···················· ···················· ···················· ···················· ····················
X 5640 hCoV-19/Scotland/QEUH-BC399A/2020|EPI_ISL_710754|2 ···················· ···················· ···················· ···················· ···················· ····················
X 5641 hCoV-19/Scotland/QEUH-BCC3DF/2020|EPI_ISL_710755|2 ···················· ···················· ···················· ···················· ···················· ····················
X 5642 hCoV-19/Scotland/QEUH-BC397C/2020|EPI_ISL_710757|2 ···················· ···················· ···················· ···················· ···················· ····················
X 5643 hCoV-19/Scotland/QEUH-BC394F/2020|EPI_ISL_710758|2 ···················· ···················· ···················· ···················· ···················· ····················
X 5644 hCoV-19/Scotland/QEUH-BCC296/2020|EPI_ISL_710759|2 ···················· ···················· ···················· ···················· ···················· ····················
X 5645 hCoV-19/Scotland/QEUH-BC38CA/2020|EPI_ISL_710760|2 ···················· ···················· ···················· ···················· ···················· ····················
X 5646 hCoV-19/Scotland/QEUH-BCC3C0/2020|EPI_ISL_710761|2 ···················· ···················· ···················· ···················· ···················· ····················
X 5647 hCoV-19/Scotland/QEUH-BCC2D2/2020|EPI_ISL_710762|2 ···················· ···················· ···················· ···················· ···················· ····················
X 5648 hCoV-19/Scotland/QEUH-BCC393/2020|EPI_ISL_710763|2 ···················· ···················· ···················· ···················· ···················· ····················
X 5649 hCoV-19/Scotland/QEUH-BCBEFD/2020|EPI_ISL_710764|2 ···················· ···················· ···················· ···················· ···················· ····················
X 5650 hCoV-19/Scotland/QEUH-BCC384/2020|EPI_ISL_710765|2 ···················· ···················· ···················· ···················· ···················· ····················
X 5651 hCoV-19/Scotland/QEUH-BC3833/2020|EPI_ISL_710766|2 ···················· ···················· ···················· ···················· ···················· ····················
X 5652 hCoV-19/Scotland/QEUH-BCC32A/2020|EPI_ISL_710770|2 ···················· ···················· ···················· ···················· ···················· ····················
X 5653 hCoV-19/Scotland/QEUH-BCC287/2020|EPI_ISL_710771|2 ···················· ···················· ···················· ···················· ···················· ····················
X 5654 hCoV-19/Scotland/QEUH-BCBEEE/2020|EPI_ISL_710778|2 ···················· ···················· ···················· ···················· ···················· ····················
X 5655 hCoV-19/Scotland/QEUH-BCC357/2020|EPI_ISL_710779|2 ···················· ···················· ···················· ···················· ···················· ····················
X 5656 hCoV-19/Scotland/QEUH-BC3903/2020|EPI_ISL_710780|2 ···················· ···················· ···················· ···················· ···················· ····················
X 5657 hCoV-19/Scotland/QEUH-BCC3FD/2020|EPI_ISL_710781|2 ···················· ···················· ···················· ···················· ···················· ····················
X 5658 hCoV-19/Scotland/QEUH-BCC409/2020|EPI_ISL_710782|2 ···················· ·A·················· ···················· ···················· ···················· ····················
X 5659 hCoV-19/Scotland/QEUH-BCC366/2020|EPI_ISL_710783|2 ···················· ···················· ···················· ···················· ···················· ····················
X 5660 hCoV-19/Scotland/QEUH-BCC348/2020|EPI_ISL_710784|2 ···················· ···················· ···················· ···················· ···················· ····················
X 5661 hCoV-19/Scotland/QEUH-BCBF63/2020|EPI_ISL_710785|2 ···················· ···················· ···················· ···················· ···················· ····················
X 5662 hCoV-19/Scotland/QEUH-BC398B/2020|EPI_ISL_710786|2 ···················· ···················· ···················· ···················· ···················· ····················
X 5663 hCoV-19/Scotland/QEUH-BC3912/2020|EPI_ISL_710787|2 ···················· ···················· ···················· ···················· ···················· ····················
X 5664 hCoV-19/Scotland/QEUH-BCBF36/2020|EPI_ISL_710788|2 ···················· ···················· ···················· ···················· ···················· ····················
X 5665 hCoV-19/Scotland/QEUH-BCC3B1/2020|EPI_ISL_710789|2 ···················· ···················· ···················· ···················· ···················· ····················
X 5666 hCoV-19/Scotland/QEUH-BCBF27/2020|EPI_ISL_710790|2 ···················· ···················· ···················· ···················· ···················· ····················
X 5667 hCoV-19/Scotland/QEUH-BC38F7/2020|EPI_ISL_710792|2 ···················· ···················· ···················· ···················· ···················· ····················
X 5668 hCoV-19/Scotland/QEUH-BCBEB1/2020|EPI_ISL_710793|2 ···················· ···················· ···················· ···················· ···················· ····················
X 5669 hCoV-19/Scotland/QEUH-BCCCB6/2020|EPI_ISL_710794|2 ···················· ···················· ···················· ···················· ···················· ····················
X 5670 hCoV-19/Scotland/QEUH-BCCB7D/2020|EPI_ISL_710795|2 ···················· ···················· ···················· ···················· ···················· ····················
X 5671 hCoV-19/Scotland/QEUH-BCCC3E/2020|EPI_ISL_710796|2 ···················· ···················· ···················· ···················· ···················· ····················
X 5672 hCoV-19/Scotland/QEUH-BCCCD4/2020|EPI_ISL_710797|2 ···················· ···················· ···················· ···················· ···················· ····················
X 5673 hCoV-19/Scotland/QEUH-BCCD59/2020|EPI_ISL_710798|2 ···················· ···················· ···················· ···················· ···················· ····················
X 5674 hCoV-19/Scotland/QEUH-BCCD77/2020|EPI_ISL_710799|2 ···················· ···················· ···················· ···················· ···················· ····················
X 5675 hCoV-19/England/CAMC-C3FA4A/2020|EPI_ISL_719113|20 ···················· ···················· ···················· ···················· ······T············· ····················
X 5676 hCoV-19/England/CAMC-C4371F/2020|EPI_ISL_719115|20 ···················· ···················· ···················· ···················· ···················· ····················
X 5677 hCoV-19/England/CAMC-C439CE/2020|EPI_ISL_719117|20 ···················· ···················· ···················· ···················· ···················· ····················
X 5678 hCoV-19/England/CAMC-C40DF2/2020|EPI_ISL_719122|20 ···················· ···················· ···················· ···················· ······T············· ····················
X 5679 hCoV-19/England/CAMC-C437F1/2020|EPI_ISL_719126|20 ···················· ···················· ···················· ···················· ···················· ····················
X 5680 hCoV-19/England/CAMC-C42B23/2020|EPI_ISL_719138|20 ···················· ···················· ···················· ···················· ···················· ····················
X 5681 hCoV-19/England/CAMC-C40B70/2020|EPI_ISL_719143|20 ···················· ···················· ···················· ···················· ···················· ····················
X 5682 hCoV-19/England/CAMC-C43AF8/2020|EPI_ISL_719148|20 ···················· ···················· ···················· ···················· ···················· ····················
X 5683 hCoV-19/England/CAMC-C4364F/2020|EPI_ISL_719151|20 ···················· ···················· ···················· ···················· ···················· ····················
X 5684 hCoV-19/England/CAMC-C41298/2020|EPI_ISL_719154|20 ···················· ···················· ···················· ···················· ···················· ····················
X 5685 hCoV-19/England/CAMC-C44EFB/2020|EPI_ISL_719160|20 ···················· ···················· ···················· ···················· ···················· ····················
X 5686 hCoV-19/England/CAMC-C424FB/2020|EPI_ISL_719163|20 ···················· ···················· ···················· ···················· ···················· ····················
X 5687 hCoV-19/England/CAMC-C43964/2020|EPI_ISL_719165|20 ···················· ···················· ···················· ···················· ···················· ····················
X 5688 hCoV-19/England/CAMC-C437C4/2020|EPI_ISL_719168|20 ···················· ···················· ···················· ···················· ···················· ····················
X 5689 hCoV-19/England/CAMC-C43B8C/2020|EPI_ISL_719180|20 ···················· ···················· ···················· ···················· ···················· ····················
X 5690 hCoV-19/England/CAMC-C439A0/2020|EPI_ISL_719182|20 ···················· ···················· ···················· ···················· ···················· ····················
X 5691 hCoV-19/England/CAMC-C422F1/2020|EPI_ISL_719188|20 ···················· ···················· ···················· ···················· ···················· ····················
X 5692 hCoV-19/England/CAMC-C43B9B/2020|EPI_ISL_719198|20 ···················· ···················· ···················· ···················· ······T············· ····················
X 5693 hCoV-19/England/MILK-BB2785/2020|EPI_ISL_720102|20 ···················· ···················· ···················· ···················· ···················· ····················
X 5694 hCoV-19/England/MILK-BB1768/2020|EPI_ISL_720103|20 ···················· ···················· ···················· ···················· ···················· ····················
X 5695 hCoV-19/England/MILK-BB256D/2020|EPI_ISL_720104|20 ···················· ···················· ···················· ···················· ···················· ····················
X 5696 hCoV-19/England/MILK-BB015C/2020|EPI_ISL_720105|20 ···················· ···················· ···················· ···················· ······T············· ····················
X 5697 hCoV-19/England/MILK-BB09CD/2020|EPI_ISL_720113|20 ···················· ·A·················· ···················· ···················· ···················· ···A················
X 5698 hCoV-19/England/MILK-BB165C/2020|EPI_ISL_720114|20 ···················· ···················· ···················· ···················· ···················· ····················
X 5699 hCoV-19/England/MILK-BB049F/2020|EPI_ISL_720115|20 ···················· ······A············· ···················· ···················· ···················· ····················
X 5700 hCoV-19/England/MILK-BB0D67/2020|EPI_ISL_720116|20 ···················· ···················· ···················· ···················· ···················· ····················

| | | | | | | | | |
|---|---|---|---|---|---|---|---|---|
| X | 5701 | hCoV-19/England/MILK-BB12D0/2020\|EPI_ISL_720117\|20 | ···················· | ···················· | ····················· | ···················· | ···················· | ···················· |
| X | 5702 | hCoV-19/England/MILK-BB0A7F/2020\|EPI_ISL_720118\|20 | ···················· | ·A·················· | ····················· | ···················· | ···················· | ··A················· |
| X | 5703 | hCoV-19/England/MILK-BB1953/2020\|EPI_ISL_720119\|20 | ···················· | ···················· | ····················· | ···················· | ···················· | ···················· |
| X | 5704 | hCoV-19/England/MILK-BB248E/2020\|EPI_ISL_720120\|20 | ···················· | ···················· | ····················· | ···················· | ···················· | ···················· |
| X | 5705 | hCoV-19/Wales/MILK-BB2020/2020\|EPI_ISL_720121\|2020 | ···················· | ···················· | ····················· | ···················· | ···················· | ···················· |
| X | 5706 | hCoV-19/England/MILK-BB1364/2020\|EPI_ISL_720122\|20 | ···················· | ···················· | ····················· | ···················· | ·····T·············· | ···················· |
| X | 5707 | hCoV-19/England/MILK-BB013E/2020\|EPI_ISL_720123\|20 | ···················· | ···················· | ····················· | ···················· | ···················· | ···················· |
| X | 5708 | hCoV-19/England/MILK-BB058D/2020\|EPI_ISL_720124\|20 | ···················· | ···················· | ····················· | ···················· | ···················· | ···················· |
| X | 5709 | hCoV-19/England/MILK-BB04CC/2020\|EPI_ISL_720125\|20 | ···················· | ···················· | ····················· | ···················· | ···················· | ···················· |
| X | 5710 | hCoV-19/England/MILK-BB0A9D/2020\|EPI_ISL_720126\|20 | ···················· | ···················· | ····················· | ···················· | ···················· | ···················· |
| X | 5711 | hCoV-19/England/MILK-BB1328/2020\|EPI_ISL_720127\|20 | ···················· | ···················· | ····················· | ···················· | ···················· | ···················· |
| X | 5712 | hCoV-19/England/MILK-BB1E63/2020\|EPI_ISL_720128\|20 | ···················· | ···················· | ····················· | ···················· | ···················· | ···················· |
| X | 5713 | hCoV-19/England/MILK-BB0FBC/2020\|EPI_ISL_720130\|20 | ···················· | ···················· | ····················· | ···················· | ···················· | ···················· |
| X | 5714 | hCoV-19/England/MILK-BB17A4/2020\|EPI_ISL_720131\|20 | ···················· | ···················· | ····················· | ···················· | ···················· | ···················· |
| X | 5715 | hCoV-19/England/MILK-BB24E8/2020\|EPI_ISL_720132\|20 | ···················· | ···················· | ····················· | ···················· | ···················· | ···················· |
| X | 5716 | hCoV-19/England/MILK-BB01B6/2020\|EPI_ISL_720134\|20 | ···················· | ···················· | ····················· | ···················· | ···················· | ···················· |
| X | 5717 | hCoV-19/England/MILK-BB2521/2020\|EPI_ISL_720135\|20 | ···················· | ···················· | ····················· | ···················· | ···················· | ···················· |
| X | 5718 | hCoV-19/England/MILK-BB18DE/2020\|EPI_ISL_720136\|20 | ···················· | ·A·················· | ····················· | ···················· | ···················· | ···················· |
| X | 5719 | hCoV-19/England/MILK-BB162F/2020\|EPI_ISL_720137\|20 | ···················· | ···················· | ····················· | ···················· | ···················· | ···················· |
| X | 5720 | hCoV-19/England/MILK-BB13FB/2020\|EPI_ISL_720138\|20 | ···················· | ···················· | ····················· | ···················· | ··A················· | ···················· |
| X | 5721 | hCoV-19/England/MILK-BB2390/2020\|EPI_ISL_720140\|20 | ···················· | ···················· | ····················· | ···················· | ···················· | ···················· |
| X | 5722 | hCoV-19/England/MILK-BB1759/2020\|EPI_ISL_720141\|20 | ···················· | ···················· | ····················· | ···················· | ···················· | ···················· |
| X | 5723 | hCoV-19/England/MILK-BB16E3/2020\|EPI_ISL_720144\|20 | ···················· | ···················· | ····················· | ···················· | ···················· | ···················· |
| X | 5724 | hCoV-19/England/MILK-BB0918/2020\|EPI_ISL_720145\|20 | ···················· | ·A·················· | ····················· | ···················· | ···················· | ··A················· |
| X | 5725 | hCoV-19/England/MILK-BB0972/2020\|EPI_ISL_720148\|20 | ···················· | ···················· | ····················· | ···················· | ···················· | ···················· |
| X | 5726 | hCoV-19/England/MILK-BB0532/2020\|EPI_ISL_720149\|20 | ···················· | ···················· | ····················· | ···················· | ···················· | ···················· |
| X | 5727 | hCoV-19/England/MILK-BB0338/2020\|EPI_ISL_720150\|20 | ···················· | ·A·················· | ····················· | ···················· | ···················· | ··A················· |
| X | 5728 | hCoV-19/England/MILK-BB1C3C/2020\|EPI_ISL_720152\|20 | ···················· | ···················· | ····················· | ···················· | ···················· | ···················· |
| X | 5729 | hCoV-19/England/MILK-BB0426/2020\|EPI_ISL_720153\|20 | ···················· | ···················· | ····················· | ···················· | ···················· | ···················· |
| X | 5730 | hCoV-19/England/MILK-BB1856/2020\|EPI_ISL_720154\|20 | ···················· | ···················· | ····················· | ···················· | ···················· | ···················· |
| X | 5731 | hCoV-19/England/MILK-BB1BD5/2020\|EPI_ISL_720155\|20 | ···················· | ·A·················· | ····················· | ···················· | ···················· | ···················· |
| X | 5732 | hCoV-19/England/MILK-BB24CA/2020\|EPI_ISL_720157\|20 | ···················· | ···················· | ····················· | ···················· | ···················· | ···················· |
| X | 5733 | hCoV-19/England/MILK-BB04EA/2020\|EPI_ISL_720158\|20 | ···················· | ···················· | ····················· | ···················· | ···················· | ···················· |
| X | 5734 | hCoV-19/England/MILK-BB104F/2020\|EPI_ISL_720159\|20 | ···················· | ···················· | ····················· | ···················· | ···················· | ···················· |
| X | 5735 | hCoV-19/England/MILK-BB14E9/2020\|EPI_ISL_720164\|20 | ···················· | ···················· | ····················· | ···················· | ···················· | ···················· |
| X | 5736 | hCoV-19/England/MILK-BB1337/2020\|EPI_ISL_720168\|20 | ···················· | ···················· | ····················· | ···················· | ···················· | ···················· |
| X | 5737 | hCoV-19/England/MILK-BB0444/2020\|EPI_ISL_720169\|20 | ···················· | ···················· | ····················· | ···················· | ···················· | ··A················· |
| X | 5738 | hCoV-19/England/MILK-BB0FDA/2020\|EPI_ISL_720171\|20 | ···················· | ···················· | ····················· | ···················· | ·····T·············· | ···················· |
| X | 5739 | hCoV-19/England/MILK-BB13BF/2020\|EPI_ISL_720174\|20 | ···················· | ···················· | ····················· | ···················· | ···················· | ···················· |
| X | 5740 | hCoV-19/England/MILK-BB15D7/2020\|EPI_ISL_720175\|20 | ···················· | ···················· | ····················· | ···················· | ···················· | ···················· |
| X | 5741 | hCoV-19/England/MILK-BB26E2/2020\|EPI_ISL_720177\|20 | ···················· | ···················· | ····················· | ···················· | ···················· | ···················· |
| X | 5742 | hCoV-19/England/MILK-BB20B7/2020\|EPI_ISL_720181\|20 | ···················· | ···················· | ····················· | ···················· | ···················· | ···················· |
| X | 5743 | hCoV-19/England/MILK-BB1926/2020\|EPI_ISL_720184\|20 | ···················· | ···················· | ····················· | ···················· | ···················· | ···················· |
| X | 5744 | hCoV-19/England/MILK-BB0FF8/2020\|EPI_ISL_720185\|20 | ···················· | ···················· | ····················· | ···················· | ···················· | ··A················· |
| X | 5745 | hCoV-19/England/MILK-BB04BD/2020\|EPI_ISL_720186\|20 | ···················· | ···················· | ····················· | ···················· | ···················· | ···················· |
| X | 5746 | hCoV-19/England/MILK-BB1540/2020\|EPI_ISL_720188\|20 | ···················· | ···················· | ····················· | ···················· | ···················· | ···················· |
| X | 5747 | hCoV-19/England/MILK-BB273A/2020\|EPI_ISL_720189\|20 | ···················· | ···················· | ····················· | ···················· | ···················· | ···················· |
| X | 5748 | hCoV-19/England/ALDP-BC6223/2020\|EPI_ISL_721400\|20 | ···················· | ···················· | ····················· | ···················· | ···················· | ···················· |
| X | 5749 | hCoV-19/England/ALDP-BC6690/2020\|EPI_ISL_721401\|20 | ···················· | ···················· | ····················· | ···················· | ···················· | ···················· |
| X | 5750 | hCoV-19/England/ALDP-BC836A/2020\|EPI_ISL_721402\|20 | ···················· | ···················· | ····················· | ···················· | ···················· | ···················· |
| X | 5751 | hCoV-19/England/ALDP-BC686D/2020\|EPI_ISL_721403\|20 | ···················· | ···················· | ····················· | ···················· | ···················· | ···················· |
| X | 5752 | hCoV-19/England/ALDP-BC7C51/2020\|EPI_ISL_721404\|20 | ···················· | ···················· | ····················· | ···················· | ···················· | ···················· |
| X | 5753 | hCoV-19/England/ALDP-BC6636/2020\|EPI_ISL_721405\|20 | ···················· | ···················· | ····················· | ···················· | ···················· | ···················· |
| X | 5754 | hCoV-19/England/ALDP-BC6487/2020\|EPI_ISL_721406\|20 | ···················· | ···················· | ····················· | ···················· | ···················· | ···················· |
| X | 5755 | hCoV-19/England/ALDP-BC8379/2020\|EPI_ISL_721407\|20 | ···················· | ···················· | ····················· | ···················· | ···················· | ···················· |
| X | 5756 | hCoV-19/England/ALDP-BC793C/2020\|EPI_ISL_721409\|20 | ···················· | ···················· | ····················· | ···················· | ···················· | ···················· |
| X | 5757 | hCoV-19/England/ALDP-BC84A3/2020\|EPI_ISL_721410\|20 | ···················· | ···················· | ····················· | ···················· | ···················· | ···················· |
| X | 5758 | hCoV-19/England/ALDP-BC8449/2020\|EPI_ISL_721411\|20 | ···················· | ···················· | ····················· | ···················· | ···················· | ···················· |
| X | 5759 | hCoV-19/England/ALDP-BC6214/2020\|EPI_ISL_721412\|20 | ···················· | ···················· | ····················· | ···················· | ···················· | ···················· |
| X | 5760 | hCoV-19/England/ALDP-BC86CB/2020\|EPI_ISL_721413\|20 | ···················· | ···················· | ····················· | ···················· | ···················· | ···················· |
| X | 5761 | hCoV-19/England/ALDP-BC7D21/2020\|EPI_ISL_721414\|20 | ···················· | ···················· | ····················· | ···················· | ···················· | ···················· |
| X | 5762 | hCoV-19/England/ALDP-BC7D9A/2020\|EPI_ISL_721415\|20 | ···················· | ···················· | ····················· | ···················· | ···················· | ···················· |
| X | 5763 | hCoV-19/England/ALDP-BC83E2/2020\|EPI_ISL_721416\|20 | ···················· | ···················· | ····················· | ···················· | ···················· | ···················· |
| X | 5764 | hCoV-19/England/ALDP-BC87F5/2020\|EPI_ISL_721417\|20 | ···················· | ···················· | ····················· | ···················· | ···················· | ···················· |
| X | 5765 | hCoV-19/England/ALDP-BC7A2A/2020\|EPI_ISL_721418\|20 | ···················· | ···················· | ····················· | ···················· | ···················· | ···················· |
| X | 5766 | hCoV-19/England/ALDP-BC7899/2020\|EPI_ISL_721419\|20 | ···················· | ···················· | ····················· | ···················· | ···················· | ···················· |
| X | 5767 | hCoV-19/England/ALDP-BC77F6/2020\|EPI_ISL_721426\|20 | ···················· | ···················· | ····················· | ···················· | ···················· | ···················· |
| X | 5768 | hCoV-19/England/ALDP-BC7802/2020\|EPI_ISL_721427\|20 | ···················· | ···················· | ····················· | ···················· | ···················· | ···················· |
| X | 5769 | hCoV-19/England/ALDP-BC7A84/2020\|EPI_ISL_721428\|20 | ···················· | ···················· | ····················· | ···················· | ···················· | ···················· |
| X | 5770 | hCoV-19/England/ALDP-BC7389/2020\|EPI_ISL_721429\|20 | ···················· | ···················· | ····················· | ···················· | ···················· | ···················· |
| X | 5771 | hCoV-19/England/ALDP-BC643C/2020\|EPI_ISL_721430\|20 | ···················· | ···················· | ····················· | ···················· | ···················· | ···················· |
| X | 5772 | hCoV-19/England/ALDP-BC7A75/2020\|EPI_ISL_721431\|20 | ···················· | ···················· | ····················· | ···················· | ···················· | ···················· |
| X | 5773 | hCoV-19/England/ALDP-BC740E/2020\|EPI_ISL_721433\|20 | ···················· | ···················· | ····················· | ···················· | ···················· | ···················· |
| X | 5774 | hCoV-19/England/ALDP-BC7969/2020\|EPI_ISL_721434\|20 | ···················· | ···················· | ····················· | ···················· | ···················· | ···················· |
| X | 5775 | hCoV-19/England/ALDP-BC78A8/2020\|EPI_ISL_721435\|20 | ···················· | ···················· | ····················· | ···················· | ···················· | ···················· |
| X | 5776 | hCoV-19/England/ALDP-BC78D5/2020\|EPI_ISL_721436\|20 | ···················· | ···················· | ····················· | ···················· | ···················· | ···················· |
| X | 5777 | hCoV-19/England/ALDP-BC7714/2020\|EPI_ISL_721437\|20 | ···················· | ···················· | ····················· | ···················· | ···················· | ···················· |
| X | 5778 | hCoV-19/England/ALDP-BC7477/2020\|EPI_ISL_721438\|20 | ···················· | ···················· | ····················· | ···················· | ···················· | ···················· |
| X | 5779 | hCoV-19/England/ALDP-BC7626/2020\|EPI_ISL_721439\|20 | ···················· | ···················· | ····················· | ···················· | ···················· | ···················· |
| X | 5780 | hCoV-19/England/ALDP-BC6232/2020\|EPI_ISL_721440\|20 | ···················· | ···················· | ····················· | ···················· | ···················· | ···················· |

| | | | | | | | |
|---|---|---|---|---|---|---|---|
| X | 5781 hCoV-19/England/ALDP-BC8458/2020\|EPI_ISL_721441\|20 | ···················· | ···················· | ···················· | ···················· | ···················· | ···················· |
| X | 5782 hCoV-19/England/ALDP-BC628D/2020\|EPI_ISL_721442\|20 | ···················· | ···················· | ···················· | ···················· | ···················· | ···················· |
| X | 5783 hCoV-19/England/ALDP-BC650C/2020\|EPI_ISL_721443\|20 | ···················· | ···················· | ···················· | ···················· | ·····T·············· | ···················· |
| X | 5784 hCoV-19/England/ALDP-BC626F/2020\|EPI_ISL_721444\|20 | ···················· | ···················· | ···················· | ···················· | ···················· | ···················· |
| X | 5785 hCoV-19/England/ALDP-BC8661/2020\|EPI_ISL_721445\|20 | ···················· | ···················· | ···················· | ···················· | ···················· | ···················· |
| X | 5786 hCoV-19/England/ALDP-BC7C24/2020\|EPI_ISL_721446\|20 | ···················· | ···················· | ···················· | ···················· | ···················· | ···················· |
| X | 5787 hCoV-19/England/ALDP-BC7D6D/2020\|EPI_ISL_721448\|20 | ···················· | ···················· | ···················· | ···················· | ···················· | ···················· |
| X | 5788 hCoV-19/England/ALDP-BC61F9/2020\|EPI_ISL_721449\|20 | ···················· | ···················· | ···················· | ···················· | ·····T·············· | ···················· |
| X | 5789 hCoV-19/England/ALDP-BC78C6/2020\|EPI_ISL_721450\|20 | ···················· | ···················· | ···················· | ···················· | ···················· | ···················· |
| X | 5790 hCoV-19/England/ALDP-BC7529/2020\|EPI_ISL_721451\|20 | ···················· | ···················· | ···················· | ···················· | ···················· | ···················· |
| X | 5791 hCoV-19/England/ALDP-BC61EA/2020\|EPI_ISL_721452\|20 | ···················· | ···················· | ···················· | ···················· | ·····T·············· | ···················· |
| X | 5792 hCoV-19/England/ALDP-BC7C8E/2020\|EPI_ISL_721454\|20 | ···················· | ···················· | ···················· | ···················· | ···················· | ···················· |
| X | 5793 hCoV-19/England/ALDP-BC7D4F/2020\|EPI_ISL_721458\|20 | ···················· | ···················· | ···················· | ···················· | ···················· | ···················· |
| X | 5794 hCoV-19/England/ALDP-BC63C6/2020\|EPI_ISL_721459\|20 | ···················· | ···················· | ···················· | ···················· | ···················· | ···················· |
| X | 5795 hCoV-19/England/ALDP-BC7BBE/2020\|EPI_ISL_721461\|20 | ···················· | ···················· | ···················· | ···················· | ···················· | ···················· |
| X | 5796 hCoV-19/England/ALDP-BC7608/2020\|EPI_ISL_721462\|20 | ···················· | ···················· | ···················· | ···················· | ···················· | ···················· |
| X | 5797 hCoV-19/England/ALDP-BC7C06/2020\|EPI_ISL_721463\|20 | ···················· | ···················· | ···················· | ···················· | ···················· | ···················· |
| X | 5798 hCoV-19/England/ALDP-BC7BEB/2020\|EPI_ISL_721465\|20 | ···················· | ···················· | ···················· | ···················· | ···················· | ···················· |
| X | 5799 hCoV-19/England/ALDP-BC8722/2020\|EPI_ISL_721466\|20 | ···················· | ···················· | ···················· | ···················· | ···················· | ···················· |
| X | 5800 hCoV-19/England/ALDP-BC8643/2020\|EPI_ISL_721467\|20 | ···················· | ···················· | ···················· | ···················· | ···················· | ···················· |
| X | 5801 hCoV-19/England/ALDP-BC87AA/2020\|EPI_ISL_721468\|20 | ···················· | ···················· | ···················· | ···················· | ···················· | ···················· |
| X | 5802 hCoV-19/England/ALDP-BC6672/2020\|EPI_ISL_721469\|20 | ···················· | ···················· | ···················· | ···················· | ···················· | ···················· |
| X | 5803 hCoV-19/England/ALDP-BC7CD9/2020\|EPI_ISL_721470\|20 | ···················· | ···················· | ···················· | ···················· | ·····T·············· | ···················· |
| X | 5804 hCoV-19/England/ALDP-BC74B3/2020\|EPI_ISL_721471\|20 | ···················· | ···················· | ···················· | ···················· | ·····T·············· | ···················· |
| X | 5805 hCoV-19/England/ALDP-BC63B7/2020\|EPI_ISL_721472\|20 | ···················· | ···················· | ···················· | ···················· | ···················· | ···················· |
| X | 5806 hCoV-19/England/ALDP-BC76CC/2020\|EPI_ISL_721474\|20 | ···················· | ···················· | ···················· | ···················· | ···················· | ···················· |
| X | 5807 hCoV-19/England/ALDP-BC79A5/2020\|EPI_ISL_721475\|20 | ···················· | ···················· | ···················· | ···················· | ···················· | ···················· |
| X | 5808 hCoV-19/England/ALDP-BC7811/2020\|EPI_ISL_721477\|20 | ···················· | ···················· | ···················· | ···················· | ···················· | ···················· |
| X | 5809 hCoV-19/England/ALDP-BC8494/2020\|EPI_ISL_721479\|20 | ···················· | ···················· | ···················· | ···················· | ···················· | ···················· |
| X | 5810 hCoV-19/England/ALDP-BC7B09/2020\|EPI_ISL_721480\|20 | ···················· | ···················· | ···················· | ···················· | ···················· | ···················· |
| X | 5811 hCoV-19/England/ALDP-BC6399/2020\|EPI_ISL_721481\|20 | ···················· | ···················· | ···················· | ···················· | ···················· | ···················· |
| X | 5812 hCoV-19/England/ALDP-BC67CA/2020\|EPI_ISL_721482\|20 | ···················· | ···················· | ···················· | ···················· | ···················· | ···················· |
| X | 5813 hCoV-19/England/ALDP-BC89B3/2020\|EPI_ISL_721483\|20 | ···················· | ·A·················· | ···················· | ···················· | ···················· | ··A················· |
| X | 5814 hCoV-19/England/ALDP-BC6162/2020\|EPI_ISL_721484\|20 | ···················· | ···················· | ···················· | ···················· | ···················· | ···················· |
| X | 5815 hCoV-19/England/ALDP-BC776F/2020\|EPI_ISL_721485\|20 | ···················· | ···················· | ···················· | ···················· | ···················· | ···················· |
| X | 5816 hCoV-19/England/ALDP-BC644B/2020\|EPI_ISL_721488\|20 | ···················· | ···················· | ···················· | ···················· | ···················· | ···················· |
| X | 5817 hCoV-19/England/ALDP-BC67E8/2020\|EPI_ISL_721489\|20 | ···················· | ···················· | ···················· | ···················· | ···················· | ···················· |
| X | 5818 hCoV-19/England/ALDP-BC73A7/2020\|EPI_ISL_721495\|20 | ···················· | ···················· | ···················· | ···················· | ···················· | ···················· |
| X | 5819 hCoV-19/England/ALDP-BC75CF/2020\|EPI_ISL_721499\|20 | ···················· | ···················· | ···················· | ···················· | ···················· | ···················· |
| X | 5820 hCoV-19/Netherlands/GE-EMC-362/2020\|EPI_ISL_722724 | ···················· | ···················· | ···················· | ···················· | ···················· | ···················· |
| X | 5821 hCoV-19/Netherlands/GE-EMC-398/2020\|EPI_ISL_722725 | ···················· | ···················· | ···················· | ···················· | ···················· | ···················· |
| X | 5822 hCoV-19/Netherlands/LI-EMC-112/2020\|EPI_ISL_722726 | ···················· | ···················· | ···················· | ···················· | ···················· | ···················· |
| X | 5823 hCoV-19/Netherlands/LI-EMC-113/2020\|EPI_ISL_722727 | ···················· | ···················· | ···················· | ···················· | ···················· | ···················· |
| X | 5824 hCoV-19/Netherlands/UT-EMC-160/2020\|EPI_ISL_722728 | ···················· | ···················· | ···················· | ···················· | ···················· | ···················· |
| X | 5825 hCoV-19/Netherlands/GE-EMC-363/2020\|EPI_ISL_722729 | ···················· | ···················· | ···················· | ···················· | ···················· | ···················· |
| X | 5826 hCoV-19/Netherlands/GE-EMC-401/2020\|EPI_ISL_722730 | ···················· | ···················· | ···················· | ···················· | ···················· | ···················· |
| X | 5827 hCoV-19/Netherlands/LI-EMC-118/2020\|EPI_ISL_722731 | ···················· | ···················· | ···················· | ···················· | ···················· | ···············A···· |
| X | 5828 hCoV-19/Netherlands/LI-EMC-119/2020\|EPI_ISL_722732 | ···················· | ···················· | ···················· | ···················· | ···················· | ···················· |
| X | 5829 hCoV-19/Netherlands/NB-EMC-415/2020\|EPI_ISL_722733 | ···················· | ···················· | ···················· | ···················· | ···················· | ···················· |
| X | 5830 hCoV-19/Netherlands/ZH-EMC-1047/2020\|EPI_ISL_72273 | ···················· | G··················· | ···················· | ···················· | ···················· | ···················· |
| X | 5831 hCoV-19/Netherlands/ZH-EMC-935/2020\|EPI_ISL_722735 | ···················· | ···················· | ···················· | ···················· | ···················· | ···················· |
| X | 5832 hCoV-19/Netherlands/ZH-EMC-986/2020\|EPI_ISL_722736 | ···················· | ···················· | ···················· | ···················· | ·····T·············· | ···················· |
| X | 5833 hCoV-19/Netherlands/GE-EMC-364/2020\|EPI_ISL_722737 | ···················· | ···················· | ···················· | ···················· | ···················· | ···················· |
| X | 5834 hCoV-19/Netherlands/LI-EMC-117/2020\|EPI_ISL_722738 | ···················· | ···················· | ···················· | ···················· | ···················· | ···················· |
| X | 5835 hCoV-19/Netherlands/LI-EMC-120/2020\|EPI_ISL_722739 | ···················· | ···················· | ···················· | ···················· | ···················· | ···················· |
| X | 5836 hCoV-19/Netherlands/UT-EMC-202/2020\|EPI_ISL_722740 | ···················· | ···················· | ···················· | ···················· | ·····T·············· | ···················· |
| X | 5837 hCoV-19/Netherlands/ZH-EMC-1030/2020\|EPI_ISL_72274 | ···················· | ···················· | ···················· | ···················· | ···················· | ···················· |
| X | 5838 hCoV-19/Netherlands/ZH-EMC-919/2020\|EPI_ISL_722742 | ···················· | ···················· | ···················· | ···················· | ···················· | ···················· |
| X | 5839 hCoV-19/Netherlands/GE-EMC-365/2020\|EPI_ISL_722743 | ···················· | ···················· | ···················· | ···················· | ···················· | ···················· |
| X | 5840 hCoV-19/Netherlands/GE-EMC-366/2020\|EPI_ISL_722744 | ···················· | ···················· | ···················· | ···················· | ···················· | ···················· |
| X | 5841 hCoV-19/Netherlands/GE-EMC-372/2020\|EPI_ISL_722745 | ···················· | ···················· | ···················· | ···················· | ···················· | ···················· |
| X | 5842 hCoV-19/Netherlands/GE-EMC-409/2020\|EPI_ISL_722746 | ···················· | ···················· | ···················· | ···················· | ···················· | ···················· |
| X | 5843 hCoV-19/Netherlands/NB-EMC-422/2020\|EPI_ISL_722747 | ···················· | ···················· | ···················· | ···················· | ···················· | ···················· |
| X | 5844 hCoV-19/Netherlands/UT-EMC-175/2020\|EPI_ISL_722748 | ···················· | ···················· | ···················· | ···················· | ···················· | ···················· |
| X | 5845 hCoV-19/Netherlands/ZH-EMC-936/2020\|EPI_ISL_722749 | ···················· | ···················· | ···················· | ···················· | ···················· | ···················· |
| X | 5846 hCoV-19/Netherlands/ZH-EMC-987/2020\|EPI_ISL_722757 | ···················· | ···················· | ···················· | ···················· | ···················· | ···················· |
| X | 5847 hCoV-19/Netherlands/GE-EMC-423/2020\|EPI_ISL_722758 | ···················· | ···················· | ···················· | ···················· | ···················· | ···················· |
| X | 5848 hCoV-19/Netherlands/GE-EMC-425/2020\|EPI_ISL_722759 | ···················· | ···················· | ···················· | ···················· | ···················· | ···················· |
| X | 5849 hCoV-19/Netherlands/GE-EMC-426/2020\|EPI_ISL_722760 | ···················· | ···················· | ···················· | ···················· | ···················· | ···················· |
| X | 5850 hCoV-19/Netherlands/GE-EMC-427/2020\|EPI_ISL_722761 | ···················· | ···················· | ···················· | ···················· | ···················· | ···················· |
| X | 5851 hCoV-19/Netherlands/GE-EMC-430/2020\|EPI_ISL_722762 | ···················· | ···················· | ···················· | ···················· | ···················· | ···················· |
| X | 5852 hCoV-19/Netherlands/UT-EMC-216/2020\|EPI_ISL_722763 | ···················· | ···················· | ···················· | ···················· | ···················· | ···················· |
| X | 5853 hCoV-19/Netherlands/ZH-EMC-1095/2020\|EPI_ISL_72276 | ···················· | ···················· | ···················· | ···················· | ···················· | ···················· |
| X | 5854 hCoV-19/Netherlands/ZH-EMC-1084/2020\|EPI_ISL_72276 | ···················· | ···················· | ···················· | ···················· | ···················· | ···················· |
| X | 5855 hCoV-19/Netherlands/ZH-EMC-1086/2020\|EPI_ISL_72276 | ···················· | ···················· | ···················· | ···················· | ···················· | ···················· |
| X | 5856 hCoV-19/Netherlands/UT-EMC-229/2020\|EPI_ISL_722767 | ···················· | ···················· | ···················· | ···················· | ···················· | ···················· |
| X | 5857 hCoV-19/Netherlands/ZH-EMC-1087/2020\|EPI_ISL_72276 | ···················· | ···················· | ···················· | ···················· | ···················· | ···················· |
| X | 5858 hCoV-19/Netherlands/ZH-EMC-1088/2020\|EPI_ISL_72276 | ···················· | G··················· | ···················· | ···················· | ···················· | ···················· |
| X | 5859 hCoV-19/Netherlands/ZH-EMC-1089/2020\|EPI_ISL_72277 | ···················· | ···················· | ···················· | ···················· | ···················· | ···················· |
| X | 5860 hCoV-19/Netherlands/ZH-EMC-1091/2020\|EPI_ISL_72277 | ···················· | ···················· | ···················· | ···················· | ···················· | ···················· |

| | | | | | | | |
|---|---|---|---|---|---|---|---|
| X | 5861 hCoV-19/Netherlands/ZH-EMC-1092/2020\|EPI_ISL_72277 | ···················· | ···················· | ····················· | ···················· | ···················· | ···················· |
| X | 5862 hCoV-19/Netherlands/ZH-EMC-1093/2020\|EPI_ISL_72277 | ···················· | ···················· | ····················· | ···················· | ···················· | ···················· |
| X | 5863 hCoV-19/Netherlands/ZH-EMC-1101/2020\|EPI_ISL_72277 | ···················· | ···················· | ····················· | ···················· | ···················· | ···················· |
| X | 5864 hCoV-19/Netherlands/ZH-EMC-1103/2020\|EPI_ISL_72277 | ···················· | ···················· | ····················· | ···················· | ···················· | ···················· |
| X | 5865 hCoV-19/Netherlands/ZH-EMC-1107/2020\|EPI_ISL_72277 | ···················· | ···················· | ····················· | ···················· | ···················· | ···················· |
| X | 5866 hCoV-19/Netherlands/ZH-EMC-1108/2020\|EPI_ISL_72277 | ···················· | ···················· | ····················· | ···················· | ···················· | ···················· |
| X | 5867 hCoV-19/Netherlands/ZH-EMC-1097/2020\|EPI_ISL_72277 | ···················· | ···················· | ····················· | ···················· | ···················· | ···················· |
| X | 5868 hCoV-19/Netherlands/ZH-EMC-1105/2020\|EPI_ISL_72277 | ···················· | ···················· | ····················· | ···················· | ···················· | ···················· |
| X | 5869 hCoV-19/Netherlands/ZH-EMC-1106/2020\|EPI_ISL_72278 | ···················· | ···················· | ····················· | ···················· | ···················· | ···················· |
| X | 5870 hCoV-19/Netherlands/ZH-EMC-1109/2020\|EPI_ISL_72278 | ···················· | ···················· | ····················· | ···················· | ···················· | ···················· |
| X | 5871 hCoV-19/Netherlands/ZH-EMC-1110/2020\|EPI_ISL_72278 | ···················· | ···················· | ····················· | ···················· | ···················· | ···················· |
| X | 5872 hCoV-19/Netherlands/ZH-EMC-1111/2020\|EPI_ISL_72278 | ···················· | ···················· | ····················· | ···················· | ···················· | ···················· |
| X | 5873 hCoV-19/Netherlands/GE-EMC-428/2020\|EPI_ISL_722784 | ···················· | ···················· | ····················· | ···················· | ···················· | ···················· |
| X | 5874 hCoV-19/Netherlands/GE-EMC-429/2020\|EPI_ISL_722785 | ···················· | ···················· | ····················· | ···················· | ···················· | ···················· |
| X | 5875 hCoV-19/Netherlands/ZH-EMC-1050/2020\|EPI_ISL_72278 | ···················· | ···················· | ····················· | ···················· | ···················· | ···················· |
| X | 5876 hCoV-19/Netherlands/ZH-EMC-1099/2020\|EPI_ISL_72278 | ···················· | ···················· | ····················· | ···················· | ···················· | ···················· |
| X | 5877 hCoV-19/Netherlands/ZH-EMC-1100/2020\|EPI_ISL_72278 | ···················· | ···················· | ····················· | ···················· | ···················· | ···················· |
| X | 5878 hCoV-19/Netherlands/UT-EMC-214/2020\|EPI_ISL_722789 | ···················· | ···················· | ····················· | ···················· | ···················· | ···················· |
| X | 5879 hCoV-19/Netherlands/UT-EMC-221/2020\|EPI_ISL_722790 | ···················· | ···················· | ····················· | ···················· | ···················· | ···················· |
| X | 5880 hCoV-19/Netherlands/ZH-EMC-1051/2020\|EPI_ISL_72279 | ···················· | ···················· | ····················· | ···················· | ···················· | ···················· |
| X | 5881 hCoV-19/Netherlands/ZH-EMC-1052/2020\|EPI_ISL_72279 | ···················· | ···················· | ····················· | ···················· | ···················· | ···················· |
| X | 5882 hCoV-19/Netherlands/ZH-EMC-1054/2020\|EPI_ISL_72279 | ···················· | ···················· | ····················· | ···················· | ···················· | ···················· |
| X | 5883 hCoV-19/Netherlands/ZH-EMC-1055/2020\|EPI_ISL_72279 | ···················· | ···················· | ····················· | ···················· | ···················· | ···················· |
| X | 5884 hCoV-19/Netherlands/ZH-EMC-1057/2020\|EPI_ISL_72279 | ···················· | ···················· | ····················· | ···················· | ···················· | ···················· |
| X | 5885 hCoV-19/Netherlands/UT-EMC-215/2020\|EPI_ISL_722796 | ···················· | ···················· | ····················· | ···················· | ···················· | ···················· |
| X | 5886 hCoV-19/Netherlands/ZH-EMC-1062/2020\|EPI_ISL_72279 | ···················· | ···················· | ····················· | ···················· | ···················· | ···················· |
| X | 5887 hCoV-19/Netherlands/ZH-EMC-1063/2020\|EPI_ISL_72279 | ···················· | G··················· | ····················· | ···················· | ···················· | ···················· |
| X | 5888 hCoV-19/Netherlands/ZH-EMC-1080/2020\|EPI_ISL_72279 | ···················· | ···················· | ····················· | ···················· | ···················· | ···················· |
| X | 5889 hCoV-19/Denmark/DCGC-13919/2020\|EPI_ISL_711301\|202 | ···················· | ···················· | ····················· | ···················· | ···················· | ···················· |
| X | 5890 hCoV-19/Denmark/DCGC-13934/2020\|EPI_ISL_711302\|202 | ···················· | ···················· | ····················· | ···················· | ···················· | ···················· |
| X | 5891 hCoV-19/Denmark/DCGC-13936/2020\|EPI_ISL_711303\|202 | ···················· | ···················· | ····················· | ···················· | ···················· | ···················· |
| X | 5892 hCoV-19/Denmark/DCGC-13943/2020\|EPI_ISL_711304\|202 | ···················· | ···················· | ····················· | ···················· | ···················· | ···················· |
| X | 5893 hCoV-19/Denmark/DCGC-13951/2020\|EPI_ISL_711305\|202 | ···················· | ···················· | ····················· | ···················· | ···················· | ···················· |
| X | 5894 hCoV-19/Denmark/DCGC-13957/2020\|EPI_ISL_711306\|202 | ···················· | ···················· | ····················· | ···················· | ···················· | ···················· |
| X | 5895 hCoV-19/Denmark/DCGC-13968/2020\|EPI_ISL_711307\|202 | ···················· | ···················· | ····················· | ···················· | ···················· | ···················· |
| X | 5896 hCoV-19/Denmark/DCGC-13979/2020\|EPI_ISL_711308\|202 | ···················· | ···················· | ····················· | ···················· | ···················· | ···················· |
| X | 5897 hCoV-19/Denmark/DCGC-13992/2020\|EPI_ISL_711309\|202 | ···················· | ···················· | ····················· | ···················· | ···················· | ···················· |
| X | 5898 hCoV-19/Denmark/DCGC-14049/2020\|EPI_ISL_711315\|202 | ···················· | ···················· | ····················· | ···················· | ···················· | ···················· |
| X | 5899 hCoV-19/Denmark/DCGC-14052/2020\|EPI_ISL_711316\|202 | ···················· | ···················· | ····················· | ···················· | ···················· | ···················· |
| X | 5900 hCoV-19/Denmark/DCGC-13127/2020\|EPI_ISL_712600\|202 | ···················· | ···················· | ····················· | ···················· | ···················· | ···················· |
| X | 5901 hCoV-19/Denmark/DCGC-13128/2020\|EPI_ISL_712601\|202 | ···················· | ···················· | ····················· | ···················· | ···················· | ···················· |
| X | 5902 hCoV-19/Denmark/DCGC-13129/2020\|EPI_ISL_712602\|202 | ···················· | ···················· | ····················· | ···················· | ···················· | ···················· |
| X | 5903 hCoV-19/Denmark/DCGC-13130/2020\|EPI_ISL_712603\|202 | ···················· | ···················· | ····················· | ···················· | ···················· | ···················· |
| X | 5904 hCoV-19/Denmark/DCGC-13162/2020\|EPI_ISL_712626\|202 | ···················· | ···················· | ····················· | ···················· | ···················· | ···················· |
| X | 5905 hCoV-19/Denmark/DCGC-13163/2020\|EPI_ISL_712627\|202 | ···················· | ···················· | ····················· | ···················· | ···················· | ···················· |
| X | 5906 hCoV-19/Denmark/DCGC-13165/2020\|EPI_ISL_712628\|202 | ···················· | ···················· | ····················· | ···················· | ···················· | ···················· |
| X | 5907 hCoV-19/Denmark/DCGC-13166/2020\|EPI_ISL_712629\|202 | ···················· | ·A·················· | ····················· | ···················· | ···················· | ··A················· |
| X | 5908 hCoV-19/Denmark/DCGC-13167/2020\|EPI_ISL_712630\|202 | ···················· | ···················· | ····················· | ···················· | ···················· | ···················· |
| X | 5909 hCoV-19/Denmark/DCGC-13168/2020\|EPI_ISL_712631\|202 | ···················· | ···················· | ····················· | ···················· | ···················· | ···················· |
| X | 5910 hCoV-19/Denmark/DCGC-13170/2020\|EPI_ISL_712632\|202 | ···················· | ···················· | ····················· | ···················· | ···················· | ···················· |
| X | 5911 hCoV-19/Denmark/DCGC-13173/2020\|EPI_ISL_712633\|202 | ···················· | ···················· | ····················· | ···················· | ···················· | ···················· |
| X | 5912 hCoV-19/Denmark/DCGC-13176/2020\|EPI_ISL_712634\|202 | ···················· | ···················· | ····················· | ···················· | ···················· | ···················· |
| X | 5913 hCoV-19/Denmark/DCGC-13180/2020\|EPI_ISL_712635\|202 | ···················· | ···················· | ····················· | ···················· | ···················· | ···················· |
| X | 5914 hCoV-19/Denmark/DCGC-13181/2020\|EPI_ISL_712636\|202 | ···················· | ···················· | ····················· | ···················· | ···················· | ···················· |
| X | 5915 hCoV-19/Denmark/DCGC-13186/2020\|EPI_ISL_712637\|202 | ···················· | ···················· | ····················· | ···················· | ···················· | ···················· |
| X | 5916 hCoV-19/Denmark/DCGC-13188/2020\|EPI_ISL_712638\|202 | ···················· | ···················· | ····················· | ···················· | ···················· | ···················· |
| X | 5917 hCoV-19/Denmark/DCGC-13189/2020\|EPI_ISL_712639\|202 | ···················· | ···················· | ····················· | ···················· | ···················· | ···················· |
| X | 5918 hCoV-19/Denmark/DCGC-13199/2020\|EPI_ISL_712646\|202 | ···················· | ···················· | ····················· | ···················· | ···················· | ···················· |
| X | 5919 hCoV-19/Denmark/DCGC-13200/2020\|EPI_ISL_712647\|202 | ···················· | ···················· | ····················· | ···················· | ···················· | ···················· |
| X | 5920 hCoV-19/Denmark/DCGC-13751/2020\|EPI_ISL_713000\|202 | ···················· | ···················· | ····················· | ···················· | ···················· | ···················· |
| X | 5921 hCoV-19/Denmark/DCGC-13752/2020\|EPI_ISL_713001\|202 | ···················· | ···················· | ····················· | ···················· | ···················· | ···················· |
| X | 5922 hCoV-19/Denmark/DCGC-13754/2020\|EPI_ISL_713002\|202 | ···················· | ···················· | ····················· | ···················· | ···················· | ···················· |
| X | 5923 hCoV-19/Denmark/DCGC-13755/2020\|EPI_ISL_713003\|202 | ···················· | ···················· | ····················· | ···················· | ···················· | ···················· |
| X | 5924 hCoV-19/Denmark/DCGC-13756/2020\|EPI_ISL_713004\|202 | ···················· | ···················· | ····················· | ···················· | ···················· | ···················· |
| X | 5925 hCoV-19/Denmark/DCGC-13758/2020\|EPI_ISL_713005\|202 | ···················· | ···················· | ····················· | ···················· | ···················· | ···················· |
| X | 5926 hCoV-19/Denmark/DCGC-13760/2020\|EPI_ISL_713006\|202 | ···················· | ·A·················· | ····················· | ···················· | ···················· | ···················· |
| X | 5927 hCoV-19/Denmark/DCGC-13763/2020\|EPI_ISL_713007\|202 | ···················· | ···················· | ····················· | ···················· | ···················· | ···················· |
| X | 5928 hCoV-19/Denmark/DCGC-13764/2020\|EPI_ISL_713008\|202 | ···················· | ·A·················· | ····················· | ···················· | ···················· | ···················· |
| X | 5929 hCoV-19/Denmark/DCGC-13765/2020\|EPI_ISL_713009\|202 | ···················· | ···················· | ····················· | ···················· | ···················· | ···················· |
| X | 5930 hCoV-19/Denmark/DCGC-13766/2020\|EPI_ISL_713010\|202 | ···················· | ···················· | ····················· | ···················· | ···················· | ···················· |
| X | 5931 hCoV-19/Denmark/DCGC-13767/2020\|EPI_ISL_713011\|202 | ···················· | ···················· | ····················· | ···················· | ···················· | ···················· |
| X | 5932 hCoV-19/Denmark/DCGC-13768/2020\|EPI_ISL_713012\|202 | ···················· | ···················· | ····················· | ···················· | ···················· | ···················· |
| X | 5933 hCoV-19/Denmark/DCGC-13769/2020\|EPI_ISL_713013\|202 | ···················· | ···················· | ····················· | ···················· | ···················· | ···················· |
| X | 5934 hCoV-19/Denmark/DCGC-13771/2020\|EPI_ISL_713014\|202 | ···················· | ···················· | ····················· | ···················· | ···················· | ···················· |
| X | 5935 hCoV-19/Denmark/DCGC-13772/2020\|EPI_ISL_713015\|202 | ···················· | ···················· | ····················· | ···················· | ···················· | ···················· |
| X | 5936 hCoV-19/Denmark/DCGC-13774/2020\|EPI_ISL_713016\|202 | ···················· | ···················· | ····················· | ···················· | ···················· | ···················· |
| X | 5937 hCoV-19/Denmark/DCGC-13776/2020\|EPI_ISL_713017\|202 | ···················· | ···················· | ····················· | ···················· | ···················· | ···················· |
| X | 5938 hCoV-19/Denmark/DCGC-13777/2020\|EPI_ISL_713018\|202 | ···················· | ···················· | ····················· | ···················· | ···················· | ···················· |
| X | 5939 hCoV-19/Denmark/DCGC-13779/2020\|EPI_ISL_713019\|202 | ···················· | ···················· | ····················· | ···················· | ···················· | ···················· |
| X | 5940 hCoV-19/Denmark/DCGC-13781/2020\|EPI_ISL_713020\|202 | ···················· | ···················· | ····················· | ···················· | ···················· | ···················· |

| | | | | | | | |
|---|---|---|---|---|---|---|---|
| X | 5941 hCoV-19/Denmark/DCGC-13782/2020\|EPI_ISL_713021\|202 | ···················· | ···················· | ···················· | ···················· | ···················· | ···················· |
| X | 5942 hCoV-19/Denmark/DCGC-13783/2020\|EPI_ISL_713022\|202 | ···················· | ···················· | ···················· | ···················· | ···················· | ···················· |
| X | 5943 hCoV-19/Denmark/DCGC-13785/2020\|EPI_ISL_713023\|202 | ···················· | ···················· | ···················· | ···················· | ···················· | ···················· |
| X | 5944 hCoV-19/Denmark/DCGC-13786/2020\|EPI_ISL_713024\|202 | ···················· | ···················· | ···················· | ···················· | ···················· | ···················· |
| X | 5945 hCoV-19/Denmark/DCGC-13787/2020\|EPI_ISL_713025\|202 | ···················· | ···················· | ···················· | ···················· | ···················· | ···················· |
| X | 5946 hCoV-19/Denmark/DCGC-13788/2020\|EPI_ISL_713026\|202 | ···················· | ···················· | ···················· | ···················· | ···················· | ···················· |
| X | 5947 hCoV-19/Denmark/DCGC-13790/2020\|EPI_ISL_713027\|202 | ···················· | ···················· | ···················· | ···················· | ···················· | ···················· |
| X | 5948 hCoV-19/Denmark/DCGC-13793/2020\|EPI_ISL_713028\|202 | ···················· | ···················· | ···················· | ···················· | ···················· | ···················· |
| X | 5949 hCoV-19/Denmark/DCGC-13795/2020\|EPI_ISL_713029\|202 | ···················· | ···················· | ···················· | ···················· | ···················· | ···················· |
| X | 5950 hCoV-19/Denmark/DCGC-13799/2020\|EPI_ISL_713030\|202 | ···················· | ···················· | ···················· | ···················· | ···················· | ···················· |
| X | 5951 hCoV-19/Denmark/DCGC-13801/2020\|EPI_ISL_713031\|202 | ···················· | ···················· | ···················· | ···················· | ···················· | ···················· |
| X | 5952 hCoV-19/Denmark/DCGC-13802/2020\|EPI_ISL_713032\|202 | ···················· | ···················· | ···················· | ···················· | ···················· | ···················· |
| X | 5953 hCoV-19/Denmark/DCGC-13803/2020\|EPI_ISL_713033\|202 | ···················· | ···················· | ···················· | ···················· | ···················· | ···················· |
| X | 5954 hCoV-19/Denmark/DCGC-13804/2020\|EPI_ISL_713034\|202 | ···················· | ···················· | ···················· | ···················· | ···················· | ···················· |
| X | 5955 hCoV-19/Denmark/DCGC-13805/2020\|EPI_ISL_713035\|202 | ···················· | ···················· | ···················· | ···················· | ···················· | ···················· |
| X | 5956 hCoV-19/Denmark/DCGC-13806/2020\|EPI_ISL_713036\|202 | ···················· | ···················· | ···················· | ···················· | ···················· | ···················· |
| X | 5957 hCoV-19/Denmark/DCGC-13808/2020\|EPI_ISL_713037\|202 | ···················· | ···················· | ···················· | ···················· | ···················· | ···················· |
| X | 5958 hCoV-19/Denmark/DCGC-13810/2020\|EPI_ISL_713038\|202 | ···················· | ···················· | ···················· | ···················· | ···················· | ···················· |
| X | 5959 hCoV-19/Denmark/DCGC-13815/2020\|EPI_ISL_713039\|202 | ···················· | ···················· | ···················· | ···················· | ···················· | ···················· |
| X | 5960 hCoV-19/Denmark/DCGC-13818/2020\|EPI_ISL_713040\|202 | ···················· | ···················· | ···················· | ···················· | ···················· | ···················· |
| X | 5961 hCoV-19/Denmark/DCGC-13819/2020\|EPI_ISL_713041\|202 | ···················· | ···················· | ···················· | ···················· | ···················· | ···················· |
| X | 5962 hCoV-19/Denmark/DCGC-13820/2020\|EPI_ISL_713042\|202 | ···················· | ···················· | ···················· | ···················· | ···················· | ···················· |
| X | 5963 hCoV-19/Denmark/DCGC-13822/2020\|EPI_ISL_713043\|202 | ···················· | ···················· | ···················· | ···················· | ···················· | ···················· |
| X | 5964 hCoV-19/Denmark/DCGC-13823/2020\|EPI_ISL_713044\|202 | ···················· | ···················· | ···················· | ···················· | ···················· | ···················· |
| X | 5965 hCoV-19/Denmark/DCGC-13824/2020\|EPI_ISL_713045\|202 | ···················· | ···················· | ···················· | ···················· | ···················· | ···················· |
| X | 5966 hCoV-19/Denmark/DCGC-13825/2020\|EPI_ISL_713046\|202 | ···················· | ···················· | ···················· | ···················· | ···················· | ···················· |
| X | 5967 hCoV-19/Denmark/DCGC-13827/2020\|EPI_ISL_713047\|202 | ···················· | ···················· | ···················· | ···················· | ···················· | ···················· |
| X | 5968 hCoV-19/Denmark/DCGC-13828/2020\|EPI_ISL_713048\|202 | ···················· | ···················· | ···················· | ···················· | ···················· | ···················· |
| X | 5969 hCoV-19/Denmark/DCGC-13830/2020\|EPI_ISL_713049\|202 | ···················· | ···················· | ···················· | ···················· | ···················· | ···················· |
| X | 5970 hCoV-19/Denmark/DCGC-13833/2020\|EPI_ISL_713050\|202 | ···················· | ···················· | ···················· | ···················· | ···················· | ···················· |
| X | 5971 hCoV-19/Denmark/DCGC-13834/2020\|EPI_ISL_713051\|202 | ···················· | ···················· | ···················· | ···················· | ···················· | ···················· |
| X | 5972 hCoV-19/Denmark/DCGC-13835/2020\|EPI_ISL_713052\|202 | ···················· | ···················· | ···················· | ···················· | ···················· | ···················· |
| X | 5973 hCoV-19/Denmark/DCGC-13837/2020\|EPI_ISL_713053\|202 | ···················· | ···················· | ···················· | ···················· | ···················· | ···················· |
| X | 5974 hCoV-19/Denmark/DCGC-13840/2020\|EPI_ISL_713054\|202 | ···················· | ···················· | ···················· | ···················· | ···················· | ···················· |
| X | 5975 hCoV-19/Denmark/DCGC-13841/2020\|EPI_ISL_713055\|202 | ···················· | ···················· | ···················· | ···················· | ···················· | ···················· |
| X | 5976 hCoV-19/Denmark/DCGC-13842/2020\|EPI_ISL_713056\|202 | ···················· | ···················· | ···················· | ···················· | ···················· | ···················· |
| X | 5977 hCoV-19/Denmark/DCGC-13843/2020\|EPI_ISL_713057\|202 | ···················· | ···················· | ···················· | ···················· | ···················· | ···················· |
| X | 5978 hCoV-19/Denmark/DCGC-13846/2020\|EPI_ISL_713058\|202 | ···················· | ···················· | ···················· | ···················· | ···················· | ···················· |
| X | 5979 hCoV-19/Denmark/DCGC-13849/2020\|EPI_ISL_713059\|202 | ···················· | ···················· | ···················· | ···················· | ···················· | ···················· |
| X | 5980 hCoV-19/Denmark/DCGC-15569/2020\|EPI_ISL_713900\|202 | ···················· | ···················· | ···················· | ···················· | ···················· | ···················· |
| X | 5981 hCoV-19/Denmark/DCGC-15570/2020\|EPI_ISL_713901\|202 | ···················· | ···················· | ···················· | ···················· | ···················· | ···················· |
| X | 5982 hCoV-19/Denmark/DCGC-15573/2020\|EPI_ISL_713902\|202 | ···················· | ···················· | ···················· | ···················· | ···················· | ···················· |
| X | 5983 hCoV-19/Denmark/DCGC-15574/2020\|EPI_ISL_713903\|202 | ···················· | ···················· | ···················· | ···················· | ···················· | ···················· |
| X | 5984 hCoV-19/Denmark/DCGC-15576/2020\|EPI_ISL_713904\|202 | ···················· | ···················· | ···················· | ···················· | ···················· | ···················· |
| X | 5985 hCoV-19/Denmark/DCGC-15578/2020\|EPI_ISL_713905\|202 | ···················· | ···················· | ···················· | ···················· | ···················· | ···················· |
| X | 5986 hCoV-19/Denmark/DCGC-15581/2020\|EPI_ISL_713906\|202 | ···················· | ···················· | ···················· | ···················· | ···················· | ···················· |
| X | 5987 hCoV-19/Denmark/DCGC-15582/2020\|EPI_ISL_713907\|202 | ···················· | ···················· | ···················· | ···················· | ···················· | ···················· |
| X | 5988 hCoV-19/Denmark/DCGC-15584/2020\|EPI_ISL_713908\|202 | ···················· | ···················· | ···················· | ···················· | ···················· | ···················· |
| X | 5989 hCoV-19/Denmark/DCGC-15585/2020\|EPI_ISL_713909\|202 | ···················· | ···················· | ···················· | ···················· | ···················· | ···················· |
| X | 5990 hCoV-19/Denmark/DCGC-15587/2020\|EPI_ISL_713910\|202 | ···················· | ···················· | ···················· | ···················· | ···················· | ···················· |
| X | 5991 hCoV-19/Denmark/DCGC-15590/2020\|EPI_ISL_713911\|202 | ···················· | ···················· | ···················· | ···················· | ···················· | ···················· |
| X | 5992 hCoV-19/Denmark/DCGC-15591/2020\|EPI_ISL_713912\|202 | ···················· | ···················· | ···················· | ···················· | ···················· | ···················· |
| X | 5993 hCoV-19/Denmark/DCGC-15592/2020\|EPI_ISL_713913\|202 | ···················· | ···················· | ···················· | ···················· | ···················· | ···················· |
| X | 5994 hCoV-19/Denmark/DCGC-15594/2020\|EPI_ISL_713914\|202 | ···················· | ···················· | ···················· | ···················· | ···················· | ···················· |
| X | 5995 hCoV-19/Denmark/DCGC-15596/2020\|EPI_ISL_713915\|202 | ···················· | ···················· | ···················· | ···················· | ···················· | ···················· |
| X | 5996 hCoV-19/Denmark/DCGC-15599/2020\|EPI_ISL_713916\|202 | ···················· | ···················· | ···················· | ···················· | ···················· | ···················· |
| X | 5997 hCoV-19/Denmark/DCGC-15601/2020\|EPI_ISL_713917\|202 | ···················· | ·A·················· | ···················· | ···················· | ···················· | ··A················· |
| X | 5998 hCoV-19/Denmark/DCGC-15604/2020\|EPI_ISL_713918\|202 | ···················· | ···················· | ···················· | ···················· | ···················· | ···················· |
| X | 5999 hCoV-19/Denmark/DCGC-15608/2020\|EPI_ISL_713919\|202 | ···················· | ···················· | ···················· | ···················· | ···················· | ···················· |
| X | 6000 hCoV-19/Denmark/DCGC-15610/2020\|EPI_ISL_713920\|202 | ···················· | ···················· | ···················· | ···················· | ···················· | ···················· |
| X | 6001 hCoV-19/Denmark/DCGC-15611/2020\|EPI_ISL_713921\|202 | ···················· | ···················· | ···················· | ···················· | ···················· | ···················· |
| X | 6002 hCoV-19/Denmark/DCGC-15612/2020\|EPI_ISL_713922\|202 | ···················· | ···················· | ···················· | ···················· | ···················· | ···················· |
| X | 6003 hCoV-19/Denmark/DCGC-15613/2020\|EPI_ISL_713923\|202 | ···················· | ···················· | ···················· | ···················· | ···················· | ···················· |
| X | 6004 hCoV-19/Denmark/DCGC-15616/2020\|EPI_ISL_713924\|202 | ···················· | ···················· | ···················· | ···················· | ···················· | ···················· |
| X | 6005 hCoV-19/Denmark/DCGC-15617/2020\|EPI_ISL_713925\|202 | ···················· | ···················· | ···················· | ···················· | ···················· | ···················· |
| X | 6006 hCoV-19/Denmark/DCGC-15619/2020\|EPI_ISL_713926\|202 | ···················· | ···················· | ···················· | ···················· | ···················· | ···················· |
| X | 6007 hCoV-19/Denmark/DCGC-15621/2020\|EPI_ISL_713927\|202 | ···················· | ···················· | ···················· | ···················· | ···················· | ···················· |
| X | 6008 hCoV-19/Denmark/DCGC-15625/2020\|EPI_ISL_713928\|202 | ···················· | ···················· | ···················· | ···················· | ···················· | ···················· |
| X | 6009 hCoV-19/Denmark/DCGC-15627/2020\|EPI_ISL_713929\|202 | ···················· | ···················· | ···················· | ···················· | ···················· | ···················· |
| X | 6010 hCoV-19/Denmark/DCGC-15628/2020\|EPI_ISL_713930\|202 | ···················· | ···················· | ···················· | ···················· | ···················· | ···················· |
| X | 6011 hCoV-19/Denmark/DCGC-15629/2020\|EPI_ISL_713931\|202 | ···················· | ···················· | ···················· | ···················· | ···················· | ···················· |
| X | 6012 hCoV-19/Denmark/DCGC-15631/2020\|EPI_ISL_713932\|202 | ···················· | ···················· | ···················· | ···················· | ···················· | ···················· |
| X | 6013 hCoV-19/Denmark/DCGC-15635/2020\|EPI_ISL_713933\|202 | ···················· | ···················· | ···················· | ···················· | ···················· | ···················· |
| X | 6014 hCoV-19/Denmark/DCGC-15637/2020\|EPI_ISL_713934\|202 | ···················· | ···················· | ···················· | ···················· | ···················· | ···················· |
| X | 6015 hCoV-19/Denmark/DCGC-15676/2020\|EPI_ISL_713957\|202 | ···················· | ···················· | ···················· | ···················· | ···················· | ···················· |
| X | 6016 hCoV-19/Denmark/DCGC-15679/2020\|EPI_ISL_713958\|202 | ···················· | ···················· | ···················· | ···················· | ···················· | ···················· |
| X | 6017 hCoV-19/Denmark/DCGC-15680/2020\|EPI_ISL_713959\|202 | ···················· | ···················· | ···················· | ···················· | ···················· | ···················· |
| X | 6018 hCoV-19/Denmark/DCGC-15681/2020\|EPI_ISL_713960\|202 | ···················· | ···················· | ···················· | ···················· | ···················· | ···················· |
| X | 6019 hCoV-19/Denmark/DCGC-15683/2020\|EPI_ISL_713961\|202 | ···················· | ···················· | ···················· | ···················· | ···················· | ···················· |
| X | 6020 hCoV-19/Denmark/DCGC-15684/2020\|EPI_ISL_713962\|202 | ···················· | ···················· | ···················· | ···················· | ···················· | ···················· |

| | | | | | | | |
|---|---|---|---|---|---|---|---|
| X | 6021 hCoV-19/Denmark/DCGC-15686/2020\|EPI_ISL_713963\|202 | ···················· | ···················· | ····················· | ···················· | ···················· | ···················· |
| X | 6022 hCoV-19/Denmark/DCGC-15687/2020\|EPI_ISL_713964\|202 | ···················· | ···················· | ····················· | ···················· | ···················· | ···················· |
| X | 6023 hCoV-19/Denmark/DCGC-15688/2020\|EPI_ISL_713965\|202 | ···················· | ···················· | ····················· | ···················· | ···················· | ···················· |
| X | 6024 hCoV-19/Denmark/DCGC-15690/2020\|EPI_ISL_713966\|202 | ···················· | ···················· | ····················· | ···················· | ···················· | ···················· |
| X | 6025 hCoV-19/Denmark/DCGC-15694/2020\|EPI_ISL_713967\|202 | ···················· | ···················· | ····················· | ···················· | ···················· | ···················· |
| X | 6026 hCoV-19/Denmark/DCGC-15695/2020\|EPI_ISL_713968\|202 | ···················· | ···················· | ····················· | ···················· | ···················· | ···················· |
| X | 6027 hCoV-19/Denmark/DCGC-15696/2020\|EPI_ISL_713969\|202 | ···················· | ···················· | ····················· | ···················· | ···················· | ···················· |
| X | 6028 hCoV-19/Denmark/DCGC-15712/2020\|EPI_ISL_713976\|202 | ···················· | ···················· | ····················· | ···················· | ···················· | ···················· |
| X | 6029 hCoV-19/Denmark/DCGC-15715/2020\|EPI_ISL_713977\|202 | ···················· | ···················· | ····················· | ···················· | ···················· | ···················· |
| X | 6030 hCoV-19/Denmark/DCGC-15716/2020\|EPI_ISL_713978\|202 | ···················· | ···················· | ····················· | ···················· | ···················· | ···················· |
| X | 6031 hCoV-19/Denmark/DCGC-15925/2020\|EPI_ISL_714300\|202 | ···················· | ···················· | ····················· | ···················· | ···················· | ···················· |
| X | 6032 hCoV-19/Denmark/DCGC-15928/2020\|EPI_ISL_714301\|202 | ···················· | ···················· | ····················· | ···················· | ···················· | ···················· |
| X | 6033 hCoV-19/Denmark/DCGC-15934/2020\|EPI_ISL_714302\|202 | ···················· | ···················· | ····················· | ···················· | ···C················ | ···················· |
| X | 6034 hCoV-19/Denmark/DCGC-15944/2020\|EPI_ISL_714303\|202 | ···················· | ···················· | ····················· | ···················· | ···················· | ···················· |
| X | 6035 hCoV-19/Denmark/DCGC-15952/2020\|EPI_ISL_714304\|202 | ···················· | ···················· | ····················· | ···················· | ···················· | ···················· |
| X | 6036 hCoV-19/Denmark/DCGC-15958/2020\|EPI_ISL_714305\|202 | ···················· | ···················· | ····················· | ···················· | ···················· | ···················· |
| X | 6037 hCoV-19/Denmark/DCGC-15964/2020\|EPI_ISL_714306\|202 | ···················· | ···················· | ····················· | ···················· | ···················· | ···················· |
| X | 6038 hCoV-19/Denmark/DCGC-15966/2020\|EPI_ISL_714307\|202 | ···················· | ···················· | ····················· | ···················· | ···················· | ···················· |
| X | 6039 hCoV-19/Denmark/DCGC-15968/2020\|EPI_ISL_714308\|202 | ···················· | ···················· | ····················· | ···················· | ···················· | ···················· |
| X | 6040 hCoV-19/Denmark/DCGC-15972/2020\|EPI_ISL_714309\|202 | ···················· | ···················· | ····················· | ···················· | ···················· | ···················· |
| X | 6041 hCoV-19/Denmark/DCGC-15977/2020\|EPI_ISL_714310\|202 | ···················· | ···················· | ····················· | ···················· | ···················· | ···················· |
| X | 6042 hCoV-19/Denmark/DCGC-15984/2020\|EPI_ISL_714311\|202 | ···················· | ···················· | ····················· | ···················· | ···················· | ···················· |
| X | 6043 hCoV-19/Denmark/DCGC-15988/2020\|EPI_ISL_714312\|202 | ···················· | ···················· | ····················· | ···················· | ···C················ | ···················· |
| X | 6044 hCoV-19/Denmark/DCGC-15992/2020\|EPI_ISL_714313\|202 | ···················· | ···················· | ····················· | ···················· | ···················· | ···················· |
| X | 6045 hCoV-19/Denmark/DCGC-16005/2020\|EPI_ISL_714314\|202 | ···················· | ···················· | ····················· | ···················· | ···················· | ···················· |
| X | 6046 hCoV-19/Denmark/DCGC-16008/2020\|EPI_ISL_714315\|202 | ···················· | ···················· | ····················· | ···················· | ···················· | ···················· |
| X | 6047 hCoV-19/Denmark/DCGC-16027/2020\|EPI_ISL_714316\|202 | ···················· | ···················· | ····················· | ···················· | ···················· | ···················· |
| X | 6048 hCoV-19/Denmark/DCGC-16035/2020\|EPI_ISL_714317\|202 | ···················· | ···················· | ····················· | ···················· | ···················· | ···················· |
| X | 6049 hCoV-19/Denmark/DCGC-16039/2020\|EPI_ISL_714318\|202 | ···················· | ···················· | ····················· | ···················· | ···················· | ···················· |
| X | 6050 hCoV-19/Denmark/DCGC-16047/2020\|EPI_ISL_714319\|202 | ···················· | ···················· | ····················· | ···················· | ···················· | ···················· |
| X | 6051 hCoV-19/Denmark/DCGC-16066/2020\|EPI_ISL_714320\|202 | ···················· | ···················· | ····················· | ···················· | ···················· | ···················· |
| X | 6052 hCoV-19/Denmark/DCGC-16073/2020\|EPI_ISL_714321\|202 | ···················· | ···················· | ····················· | ···················· | ···················· | ···················· |
| X | 6053 hCoV-19/Denmark/DCGC-16079/2020\|EPI_ISL_714322\|202 | ···················· | ···················· | ····················· | ···················· | ···················· | ···················· |
| X | 6054 hCoV-19/Denmark/DCGC-16081/2020\|EPI_ISL_714323\|202 | ···················· | ···················· | ····················· | ···················· | ···················· | ···················· |
| X | 6055 hCoV-19/Denmark/DCGC-16102/2020\|EPI_ISL_714324\|202 | ···················· | ···················· | ····················· | ···················· | ···················· | ···················· |
| X | 6056 hCoV-19/Denmark/DCGC-16126/2020\|EPI_ISL_714325\|202 | ···················· | ···················· | ····················· | ···················· | ···················· | ···················· |
| X | 6057 hCoV-19/Denmark/DCGC-16169/2020\|EPI_ISL_714326\|202 | ···················· | ···················· | ····················· | ···················· | ···················· | ···················· |
| X | 6058 hCoV-19/Denmark/DCGC-16171/2020\|EPI_ISL_714327\|202 | ···················· | ···················· | ····················· | ···················· | ···················· | ···················· |
| X | 6059 hCoV-19/Denmark/DCGC-16176/2020\|EPI_ISL_714328\|202 | ···················· | ···················· | ····················· | ···················· | ···················· | ···················· |
| X | 6060 hCoV-19/Denmark/DCGC-16181/2020\|EPI_ISL_714329\|202 | ···················· | ···················· | ····················· | ···················· | ···················· | ···················· |
| X | 6061 hCoV-19/Denmark/DCGC-16186/2020\|EPI_ISL_714330\|202 | ···················· | ···················· | ····················· | ···················· | ···················· | ···················· |
| X | 6062 hCoV-19/Denmark/DCGC-16192/2020\|EPI_ISL_714331\|202 | ···················· | ···················· | ····················· | ···················· | ···················· | ···················· |
| X | 6063 hCoV-19/Denmark/DCGC-16198/2020\|EPI_ISL_714332\|202 | ···················· | ···················· | ····················· | ···················· | ···················· | ···················· |
| X | 6064 hCoV-19/Denmark/DCGC-16212/2020\|EPI_ISL_714333\|202 | ···················· | ···················· | ····················· | ···················· | ···················· | ···················· |
| X | 6065 hCoV-19/Denmark/DCGC-16215/2020\|EPI_ISL_714334\|202 | ···················· | ···················· | ····················· | ···················· | ···················· | ···················· |
| X | 6066 hCoV-19/Denmark/DCGC-16217/2020\|EPI_ISL_714335\|202 | ···················· | ···················· | ····················· | ···················· | ···················· | ···················· |
| X | 6067 hCoV-19/Denmark/DCGC-16222/2020\|EPI_ISL_714336\|202 | ···················· | ···················· | ····················· | ···················· | ···················· | ···················· |
| X | 6068 hCoV-19/Denmark/DCGC-16244/2020\|EPI_ISL_714337\|202 | ···················· | ···················· | ····················· | ···················· | ···················· | ···················· |
| X | 6069 hCoV-19/Denmark/DCGC-16279/2020\|EPI_ISL_714338\|202 | ···················· | ···················· | ····················· | ···················· | ···················· | ···················· |
| X | 6070 hCoV-19/Denmark/DCGC-16311/2020\|EPI_ISL_714339\|202 | ···················· | ···················· | ····················· | ···················· | ···················· | ···················· |
| X | 6071 hCoV-19/Denmark/DCGC-16319/2020\|EPI_ISL_714340\|202 | ···················· | ···················· | ····················· | ···················· | ···················· | ···················· |
| X | 6072 hCoV-19/Denmark/DCGC-16329/2020\|EPI_ISL_714341\|202 | ···················· | ···················· | ····················· | ···················· | ···················· | ···················· |
| X | 6073 hCoV-19/Denmark/DCGC-16350/2020\|EPI_ISL_714342\|202 | ···················· | ···················· | ····················· | ···················· | ···················· | ···················· |
| X | 6074 hCoV-19/Denmark/DCGC-16352/2020\|EPI_ISL_714343\|202 | ···················· | ···················· | ····················· | ···················· | ···················· | ···················· |
| X | 6075 hCoV-19/Denmark/DCGC-16355/2020\|EPI_ISL_714344\|202 | ···················· | ···················· | ····················· | ···················· | ···················· | ···················· |
| X | 6076 hCoV-19/Denmark/DCGC-16374/2020\|EPI_ISL_714345\|202 | ···················· | ···················· | ····················· | ···················· | ···················· | ···················· |
| X | 6077 hCoV-19/Denmark/DCGC-16381/2020\|EPI_ISL_714346\|202 | ···················· | ···················· | ····················· | ···················· | ···················· | ···················· |
| X | 6078 hCoV-19/Denmark/DCGC-16451/2020\|EPI_ISL_714347\|202 | ···················· | ···················· | ····················· | ···················· | ···················· | ···················· |
| X | 6079 hCoV-19/Denmark/DCGC-16494/2020\|EPI_ISL_714348\|202 | ···················· | ···················· | ····················· | ···················· | ···················· | ···················· |
| X | 6080 hCoV-19/Denmark/DCGC-16497/2020\|EPI_ISL_714349\|202 | ···················· | ···················· | ····················· | ···················· | ···················· | ···················· |
| X | 6081 hCoV-19/Denmark/DCGC-16522/2020\|EPI_ISL_714350\|202 | ···················· | ···················· | ····················· | ···················· | ···················· | ···················· |
| X | 6082 hCoV-19/Denmark/DCGC-16539/2020\|EPI_ISL_714351\|202 | ···················· | ···················· | ····················· | ···················· | ···················· | ···················· |
| X | 6083 hCoV-19/Denmark/DCGC-16549/2020\|EPI_ISL_714352\|202 | ···················· | ···················· | ····················· | ···················· | ···················· | ···················· |
| X | 6084 hCoV-19/Denmark/DCGC-16564/2020\|EPI_ISL_714353\|202 | ···················· | ···················· | ····················· | ···················· | ···C················ | ···················· |
| X | 6085 hCoV-19/Denmark/DCGC-16573/2020\|EPI_ISL_714354\|202 | ···················· | ···················· | ····················· | ···················· | ···················· | ···················· |
| X | 6086 hCoV-19/Denmark/DCGC-16575/2020\|EPI_ISL_714355\|202 | ···················· | ···················· | ····················· | ···················· | ···················· | ···················· |
| X | 6087 hCoV-19/Denmark/DCGC-16591/2020\|EPI_ISL_714356\|202 | ···················· | ···················· | ····················· | ···················· | ···················· | ···················· |
| X | 6088 hCoV-19/Denmark/DCGC-16596/2020\|EPI_ISL_714357\|202 | ···················· | ···················· | ····················· | ···················· | ···················· | ···················· |
| X | 6089 hCoV-19/Denmark/DCGC-16618/2020\|EPI_ISL_714358\|202 | ···················· | ···················· | ····················· | ···················· | ···················· | ···················· |
| X | 6090 hCoV-19/Denmark/DCGC-16624/2020\|EPI_ISL_714359\|202 | ···················· | ···················· | ····················· | ···················· | ···················· | ···················· |
| X | 6091 hCoV-19/Denmark/DCGC-16625/2020\|EPI_ISL_714360\|202 | ···················· | ···················· | ····················· | ···················· | ···················· | ···················· |
| X | 6092 hCoV-19/Denmark/DCGC-16628/2020\|EPI_ISL_714361\|202 | ···················· | ···················· | ····················· | ···················· | ···················· | ···················· |
| X | 6093 hCoV-19/Denmark/DCGC-16644/2020\|EPI_ISL_714362\|202 | ···················· | ···················· | ····················· | ···················· | ···················· | ···················· |
| X | 6094 hCoV-19/Denmark/DCGC-16668/2020\|EPI_ISL_714363\|202 | ···················· | ···················· | ····················· | ···················· | ···················· | ···················· |
| X | 6095 hCoV-19/Denmark/DCGC-16735/2020\|EPI_ISL_714364\|202 | ···················· | ···················· | ····················· | ···················· | ···················· | ···················· |
| X | 6096 hCoV-19/Denmark/DCGC-15981/2020\|EPI_ISL_714366\|202 | ···················· | ·A·················· | ····················· | ···················· | ···················· | ···················· |
| X | 6097 hCoV-19/Denmark/DCGC-16204/2020\|EPI_ISL_714367\|202 | ···················· | ·N·················· | ····················· | ···················· | ···················· | ···················· |
| X | 6098 hCoV-19/Denmark/DCGC-16132/2020\|EPI_ISL_714372\|202 | ···················· | ·A·················· | ····················· | ···················· | ···················· | ···A················ |
| X | 6099 hCoV-19/Denmark/DCGC-16145/2020\|EPI_ISL_714373\|202 | ···················· | ·A·················· | ····················· | ···················· | ···················· | ···A················ |
| X | 6100 hCoV-19/Denmark/DCGC-16252/2020\|EPI_ISL_714374\|202 | ···················· | ···················· | ····················· | ···················· | ···················· | ···················· |

| | | | | | | | |
|---|---|---|---|---|---|---|---|
| X | 6101 hCoV-19/Denmark/DCGC-16295/2020\|EPI_ISL_714375\|202 | ···················· | ···················· | ····················· | ···················· | ···················· | ···················· |
| X | 6102 hCoV-19/Denmark/DCGC-16610/2020\|EPI_ISL_714376\|202 | ···················· | ···················· | ····················· | ···················· | ·····T·············· | ···················· |
| X | 6103 hCoV-19/Denmark/DCGC-16697/2020\|EPI_ISL_714377\|202 | ···················· | ···················· | ····················· | ···················· | ···················· | ···················· |
| X | 6104 hCoV-19/Denmark/DCGC-16139/2020\|EPI_ISL_714379\|202 | ···················· | ·A·················· | ····················· | ···················· | ···················· | ···A················ |
| X | 6105 hCoV-19/Denmark/DCGC-16361/2020\|EPI_ISL_714380\|202 | ···················· | ···················· | ····················· | ···················· | ···················· | ···················· |
| X | 6106 hCoV-19/England/ALDP-C481D0/2020\|EPI_ISL_718750\|20 | ···················· | ···················· | ····················· | ···················· | ···················· | ···················· |
| X | 6107 hCoV-19/England/ALDP-C47F22/2020\|EPI_ISL_718754\|20 | ···················· | ···················· | ····················· | ···················· | ···················· | ···················· |
| X | 6108 hCoV-19/England/CAMC-C46EBD/2020\|EPI_ISL_718756\|20 | ···················· | ···················· | ····················· | ···················· | ···················· | ···················· |
| X | 6109 hCoV-19/England/ALDP-C49C1A/2020\|EPI_ISL_718759\|20 | ···················· | ···················· | ····················· | ···················· | ···················· | ···················· |
| X | 6110 hCoV-19/England/ALDP-C497FB/2020\|EPI_ISL_718761\|20 | ···················· | ···················· | ····················· | ···················· | ···················· | ···················· |
| X | 6111 hCoV-19/England/ALDP-C47E61/2020\|EPI_ISL_718762\|20 | ···················· | ···················· | ····················· | ···················· | ···················· | ···················· |
| X | 6112 hCoV-19/England/ALDP-C498AD/2020\|EPI_ISL_718764\|20 | ···················· | ···················· | ····················· | ···················· | ···················· | ···················· |
| X | 6113 hCoV-19/England/ALDP-C4803D/2020\|EPI_ISL_718766\|20 | ···················· | ·A·················· | ····················· | ···················· | ···················· | ···A················ |
| X | 6114 hCoV-19/England/ALDP-C481EF/2020\|EPI_ISL_718767\|20 | ···················· | ···················· | ····················· | ···················· | ···················· | ···················· |
| X | 6115 hCoV-19/England/ALDP-C47DBF/2020\|EPI_ISL_718769\|20 | ···················· | ···················· | ····················· | ···················· | ···················· | ···················· |
| X | 6116 hCoV-19/England/ALDP-C49834/2020\|EPI_ISL_718772\|20 | ···················· | ···················· | ····················· | ···················· | ···················· | ···················· |
| X | 6117 hCoV-19/England/ALDP-C47E8F/2020\|EPI_ISL_718774\|20 | ···················· | ···················· | ····················· | ···················· | ···················· | ···A················ |
| X | 6118 hCoV-19/England/ALDP-C48149/2020\|EPI_ISL_718777\|20 | ···················· | ···················· | ····················· | ···················· | ···················· | ···················· |
| X | 6119 hCoV-19/England/ALDP-C47C76/2020\|EPI_ISL_718779\|20 | ···················· | ···················· | ····················· | ···················· | ···················· | ···················· |
| X | 6120 hCoV-19/England/CAMC-C4756C/2020\|EPI_ISL_718785\|20 | ···················· | ···················· | ····················· | ···················· | ···················· | ···················· |
| X | 6121 hCoV-19/England/CAMC-C47229/2020\|EPI_ISL_718788\|20 | ···················· | ···················· | ····················· | ···················· | ···················· | ···················· |
| X | 6122 hCoV-19/England/ALDP-C4820A/2020\|EPI_ISL_718791\|20 | ···················· | ···················· | ····················· | ···················· | ···················· | ···················· |
| X | 6123 hCoV-19/England/ALDP-C49A5C/2020\|EPI_ISL_718795\|20 | ···················· | ···················· | ····················· | ···················· | ···················· | ···················· |
| X | 6124 hCoV-19/England/CAMC-C44F25/2020\|EPI_ISL_719173\|20 | ···················· | ···················· | ····················· | ···················· | ···················· | ···················· |
| X | 6125 hCoV-19/England/CAMC-C44ECE/2020\|EPI_ISL_719176\|20 | ···················· | ···················· | ····················· | ···················· | ·····T·············· | ···················· |
| X | 6126 hCoV-19/England/MILK-BB059C/2020\|EPI_ISL_720161\|20 | ···················· | ···················· | ····················· | ···················· | ···················· | ···················· |
| X | 6127 hCoV-19/England/MILK-BB270D/2020\|EPI_ISL_720162\|20 | ···················· | ···················· | ····················· | ···················· | ···················· | ···················· |
| X | 6128 hCoV-19/England/MILK-BB00C8/2020\|EPI_ISL_720165\|20 | ···················· | ···················· | ····················· | ···················· | ···················· | ···················· |
| X | 6129 hCoV-19/England/MILK-BB249D/2020\|EPI_ISL_720166\|20 | ···················· | ···················· | ····················· | ···················· | ···················· | ···················· |
| X | 6130 hCoV-19/England/MILK-BB12C1/2020\|EPI_ISL_720167\|20 | ···················· | ···················· | ····················· | ···················· | ···················· | ···················· |
| X | 6131 hCoV-19/England/MILK-BB257C/2020\|EPI_ISL_720191\|20 | ···················· | ···················· | ····················· | ···················· | ···················· | ···················· |
| X | 6132 hCoV-19/England/MILK-BB1EBE/2020\|EPI_ISL_720192\|20 | ···················· | ·A·················· | ····················· | ···················· | ···················· | ···A················ |
| X | 6133 hCoV-19/England/MILK-BB254F/2020\|EPI_ISL_720194\|20 | ···················· | ···················· | ····················· | ···················· | ···················· | ···················· |
| X | 6134 hCoV-19/England/MILK-BB1698/2020\|EPI_ISL_720195\|20 | ···················· | ···················· | ····················· | ···················· | ···················· | ···················· |
| X | 6135 hCoV-19/England/MILK-BB0936/2020\|EPI_ISL_720197\|20 | ···················· | ···················· | ····················· | ···················· | ···················· | ···················· |
| X | 6136 hCoV-19/England/MILK-BB123A/2020\|EPI_ISL_720198\|20 | ···················· | ···················· | ····················· | ···················· | ···················· | ···················· |
| X | 6137 hCoV-19/England/ALDP-BC8625/2020\|EPI_ISL_721490\|20 | ···················· | ···················· | ····················· | ···················· | ···················· | ···················· |
| X | 6138 hCoV-19/England/ALDP-BC7A0C/2020\|EPI_ISL_721491\|20 | ···················· | ···················· | ····················· | ···················· | ···················· | ···················· |
| X | 6139 hCoV-19/England/ALDP-BC8476/2020\|EPI_ISL_721492\|20 | ···················· | ···················· | ····················· | ···················· | ···················· | ···················· |
| X | 6140 hCoV-19/England/ALDP-BC67F7/2020\|EPI_ISL_721493\|20 | ···················· | ···················· | ····················· | ···················· | ···················· | ···················· |
| X | 6141 hCoV-19/England/ALDP-BC77BA/2020\|EPI_ISL_721494\|20 | ···················· | ···················· | ····················· | ···················· | ···················· | ···················· |
| X | 6142 hCoV-19/England/ALDP-BC7723/2020\|EPI_ISL_721496\|20 | ···················· | ···················· | ····················· | ···················· | ···················· | ···················· |
| X | 6143 hCoV-19/England/ALDP-BC7B81/2020\|EPI_ISL_721497\|20 | ···················· | ···················· | ····················· | ···················· | ···················· | ···················· |
| X | 6144 hCoV-19/England/ALDP-BC8467/2020\|EPI_ISL_721498\|20 | ···················· | ···················· | ····················· | ···················· | ···················· | ···················· |
| X | 6145 hCoV-19/Denmark/DCGC-13994/2020\|EPI_ISL_711310\|202 | ···················· | ···················· | ····················· | ···················· | ···················· | ···················· |
| X | 6146 hCoV-19/Denmark/DCGC-14032/2020\|EPI_ISL_711311\|202 | ···················· | ···················· | ····················· | ···················· | ···················· | ···················· |
| X | 6147 hCoV-19/Denmark/DCGC-14033/2020\|EPI_ISL_711312\|202 | ···················· | ···················· | ····················· | ···················· | ···················· | ···················· |
| X | 6148 hCoV-19/Denmark/DCGC-14042/2020\|EPI_ISL_711313\|202 | ···················· | ···················· | ····················· | ···················· | ···················· | ···················· |
| X | 6149 hCoV-19/Denmark/DCGC-14047/2020\|EPI_ISL_711314\|202 | ···················· | ···················· | ····················· | ···················· | ···················· | ···················· |
| X | 6150 hCoV-19/Denmark/DCGC-14070/2020\|EPI_ISL_711317\|202 | ···················· | ···················· | ····················· | ···················· | ···················· | ···················· |
| X | 6151 hCoV-19/Denmark/DCGC-14078/2020\|EPI_ISL_711318\|202 | ···················· | ···················· | ····················· | ···················· | ···················· | ···················· |
| X | 6152 hCoV-19/Denmark/DCGC-14084/2020\|EPI_ISL_711319\|202 | ···················· | ···················· | ····················· | ···················· | ···················· | ···················· |
| X | 6153 hCoV-19/Denmark/DCGC-14086/2020\|EPI_ISL_711320\|202 | ···················· | ···················· | ····················· | ···················· | ···················· | ···················· |
| X | 6154 hCoV-19/Denmark/DCGC-14100/2020\|EPI_ISL_711321\|202 | ···················· | ···················· | ····················· | ···················· | ···················· | ···················· |
| X | 6155 hCoV-19/Denmark/DCGC-14124/2020\|EPI_ISL_711322\|202 | ···················· | ···················· | ····················· | ···················· | ···················· | ···················· |
| X | 6156 hCoV-19/Denmark/DCGC-14128/2020\|EPI_ISL_711323\|202 | ···················· | ···················· | ····················· | ···················· | ···C················ | ···················· |
| X | 6157 hCoV-19/Denmark/DCGC-14140/2020\|EPI_ISL_711324\|202 | ···················· | ···················· | ····················· | ···················· | ···················· | ···················· |
| X | 6158 hCoV-19/Denmark/DCGC-14151/2020\|EPI_ISL_711325\|202 | ···················· | ···················· | ····················· | ···················· | ···C················ | ···················· |
| X | 6159 hCoV-19/Denmark/DCGC-14155/2020\|EPI_ISL_711326\|202 | ···················· | ···················· | ····················· | ···················· | ···················· | ···················· |
| X | 6160 hCoV-19/Denmark/DCGC-14165/2020\|EPI_ISL_711327\|202 | ···················· | ···················· | ····················· | ···················· | ···················· | ···················· |
| X | 6161 hCoV-19/Denmark/DCGC-14172/2020\|EPI_ISL_711328\|202 | ···················· | ···················· | ····················· | ···················· | ···················· | ···················· |
| X | 6162 hCoV-19/Denmark/DCGC-14184/2020\|EPI_ISL_711329\|202 | ···················· | ···················· | ····················· | ···················· | ···················· | ···················· |
| X | 6163 hCoV-19/Denmark/DCGC-14187/2020\|EPI_ISL_711330\|202 | ···················· | ···················· | ····················· | ···················· | ···················· | ···················· |
| X | 6164 hCoV-19/Denmark/DCGC-14190/2020\|EPI_ISL_711331\|202 | ···················· | ···················· | ····················· | ···················· | ···················· | ···················· |
| X | 6165 hCoV-19/Denmark/DCGC-14191/2020\|EPI_ISL_711332\|202 | ···················· | ···················· | ····················· | ···················· | ···················· | ···················· |
| X | 6166 hCoV-19/Denmark/DCGC-14196/2020\|EPI_ISL_711333\|202 | ···················· | ···················· | ····················· | ···················· | ···C················ | ···················· |
| X | 6167 hCoV-19/Denmark/DCGC-14211/2020\|EPI_ISL_711334\|202 | ···················· | ···················· | ····················· | ···················· | ···················· | ···················· |
| X | 6168 hCoV-19/Denmark/DCGC-14223/2020\|EPI_ISL_711335\|202 | ···················· | ···················· | ····················· | ···················· | ···C················ | ···················· |
| X | 6169 hCoV-19/Denmark/DCGC-14251/2020\|EPI_ISL_711336\|202 | ···················· | ···················· | ····················· | ···················· | ···················· | ···················· |
| X | 6170 hCoV-19/Denmark/DCGC-14258/2020\|EPI_ISL_711337\|202 | ···················· | ···················· | ····················· | ···················· | ···················· | ···················· |
| X | 6171 hCoV-19/Denmark/DCGC-14259/2020\|EPI_ISL_711338\|202 | ···················· | ···················· | ····················· | ···················· | ···················· | ···················· |
| X | 6172 hCoV-19/Denmark/DCGC-14262/2020\|EPI_ISL_711339\|202 | ···················· | ···················· | ····················· | ···················· | ···················· | ···················· |
| X | 6173 hCoV-19/Denmark/DCGC-14274/2020\|EPI_ISL_711340\|202 | ···················· | ···················· | ····················· | ···················· | ···················· | ···················· |
| X | 6174 hCoV-19/Denmark/DCGC-14277/2020\|EPI_ISL_711341\|202 | ···················· | ···················· | ····················· | ···················· | ···················· | ···················· |
| X | 6175 hCoV-19/Denmark/DCGC-14278/2020\|EPI_ISL_711342\|202 | ···················· | ···················· | ····················· | ···················· | ···················· | ···················· |
| X | 6176 hCoV-19/Denmark/DCGC-14286/2020\|EPI_ISL_711343\|202 | ···················· | ···················· | ····················· | ···················· | ···················· | ···················· |
| X | 6177 hCoV-19/Denmark/DCGC-14289/2020\|EPI_ISL_711344\|202 | ···················· | ···················· | ····················· | ···················· | ···················· | ···················· |
| X | 6178 hCoV-19/Denmark/DCGC-14316/2020\|EPI_ISL_711345\|202 | ···················· | ···················· | ····················· | ···················· | ···················· | ···················· |
| X | 6179 hCoV-19/Denmark/DCGC-14320/2020\|EPI_ISL_711346\|202 | ···················· | ···················· | ····················· | ···················· | ···················· | ···················· |
| X | 6180 hCoV-19/Denmark/DCGC-14325/2020\|EPI_ISL_711347\|202 | ···················· | ···················· | ····················· | ···················· | ···················· | ···················· |

| | | | | | | | | |
|---|---|---|---|---|---|---|---|---|
| X | 6181 | hCoV-19/Denmark/DCGC-14353/2020\|EPI_ISL_711348\|202 | .................... | .................... | .................... | .................... | .................... | .................... |
| X | 6182 | hCoV-19/Denmark/DCGC-14360/2020\|EPI_ISL_711349\|202 | .................... | .................... | .................... | .................... | .................... | .................... |
| X | 6183 | hCoV-19/Denmark/DCGC-14409/2020\|EPI_ISL_711350\|202 | .................... | .................... | .................... | .................... | .................... | .................... |
| X | 6184 | hCoV-19/Denmark/DCGC-14429/2020\|EPI_ISL_711351\|202 | .................... | .................... | .................... | .................... | ...C................ | .................... |
| X | 6185 | hCoV-19/Denmark/DCGC-14431/2020\|EPI_ISL_711352\|202 | .................... | .................... | .................... | .................... | .................... | .................... |
| X | 6186 | hCoV-19/Denmark/DCGC-14452/2020\|EPI_ISL_711353\|202 | .................... | .................... | .................... | .................... | .................... | .................... |
| X | 6187 | hCoV-19/Denmark/DCGC-14467/2020\|EPI_ISL_711354\|202 | .................... | .................... | .................... | .................... | .................... | .................... |
| X | 6188 | hCoV-19/Denmark/DCGC-14482/2020\|EPI_ISL_711355\|202 | .................... | .................... | .................... | .................... | .................... | .................... |
| X | 6189 | hCoV-19/Denmark/DCGC-14487/2020\|EPI_ISL_711356\|202 | .................... | .................... | .................... | .................... | .................... | .................... |
| X | 6190 | hCoV-19/Denmark/DCGC-14500/2020\|EPI_ISL_711357\|202 | .................... | .................... | .................... | .................... | .................... | .................... |
| X | 6191 | hCoV-19/Denmark/DCGC-14521/2020\|EPI_ISL_711358\|202 | .................... | .................... | .................... | .................... | .................... | .................... |
| X | 6192 | hCoV-19/Denmark/DCGC-14524/2020\|EPI_ISL_711359\|202 | .................... | .................... | .................... | .................... | .................... | .................... |
| X | 6193 | hCoV-19/Denmark/DCGC-14626/2020\|EPI_ISL_711364\|202 | .................... | .................... | .................... | .................... | .................... | .................... |
| X | 6194 | hCoV-19/Denmark/DCGC-14640/2020\|EPI_ISL_711365\|202 | .................... | .................... | .................... | .................... | .................... | .................... |
| X | 6195 | hCoV-19/Denmark/DCGC-14641/2020\|EPI_ISL_711366\|202 | .................... | .................... | .................... | .................... | .................... | .................... |
| X | 6196 | hCoV-19/Denmark/DCGC-14647/2020\|EPI_ISL_711367\|202 | .................... | .................... | .................... | .................... | .................... | .................... |
| X | 6197 | hCoV-19/Denmark/DCGC-14756/2020\|EPI_ISL_711368\|202 | .................... | .................... | .................... | .................... | .................... | .................... |
| X | 6198 | hCoV-19/Denmark/DCGC-14763/2020\|EPI_ISL_711369\|202 | .................... | .................... | .................... | .................... | .................... | .................... |
| X | 6199 | hCoV-19/Denmark/DCGC-14884/2020\|EPI_ISL_711380\|202 | .................... | .................... | .................... | .................... | .................... | .................... |
| X | 6200 | hCoV-19/Denmark/DCGC-14886/2020\|EPI_ISL_711381\|202 | .................... | .................... | .................... | .................... | .................... | .................... |
| X | 6201 | hCoV-19/Denmark/DCGC-14900/2020\|EPI_ISL_711382\|202 | .................... | .................... | .................... | .................... | .................... | .................... |
| X | 6202 | hCoV-19/Denmark/DCGC-14901/2020\|EPI_ISL_711383\|202 | .................... | .................... | .................... | .................... | .................... | .................... |
| X | 6203 | hCoV-19/Denmark/DCGC-14009/2020\|EPI_ISL_711388\|202 | .................... | .................... | .................... | .................... | .................... | .................... |
| X | 6204 | hCoV-19/Denmark/DCGC-14116/2020\|EPI_ISL_711389\|202 | .................... | .................... | .................... | .................... | .................... | .................... |
| X | 6205 | hCoV-19/Denmark/DCGC-14903/2020\|EPI_ISL_711390\|202 | .................... | .................... | .................... | .................... | .................... | .................... |
| X | 6206 | hCoV-19/Denmark/DCGC-14346/2020\|EPI_ISL_711394\|202 | .................... | .A.................. | .................... | .................... | .................... | ...A................ |
| X | 6207 | hCoV-19/Denmark/DCGC-13132/2020\|EPI_ISL_712604\|202 | .................... | .................... | .................... | .................... | .................... | .................... |
| X | 6208 | hCoV-19/Denmark/DCGC-13133/2020\|EPI_ISL_712605\|202 | .................... | .................... | .................... | .................... | .................... | .................... |
| X | 6209 | hCoV-19/Denmark/DCGC-13135/2020\|EPI_ISL_712606\|202 | .................... | .................... | .................... | .................... | .................... | .................... |
| X | 6210 | hCoV-19/Denmark/DCGC-13136/2020\|EPI_ISL_712607\|202 | .................... | .................... | .................... | .................... | .................... | .................... |
| X | 6211 | hCoV-19/Denmark/DCGC-13138/2020\|EPI_ISL_712608\|202 | .................... | .................... | .................... | .................... | .................... | .................... |
| X | 6212 | hCoV-19/Denmark/DCGC-13139/2020\|EPI_ISL_712609\|202 | .................... | .................... | .................... | .................... | .................... | .................... |
| X | 6213 | hCoV-19/Denmark/DCGC-13140/2020\|EPI_ISL_712610\|202 | .................... | .................... | .................... | .................... | .................... | .................... |
| X | 6214 | hCoV-19/Denmark/DCGC-13141/2020\|EPI_ISL_712611\|202 | .................... | .A.................. | .................... | .................... | .................... | .................... |
| X | 6215 | hCoV-19/Denmark/DCGC-13142/2020\|EPI_ISL_712612\|202 | .................... | .................... | .................... | .................... | .................... | .................... |
| X | 6216 | hCoV-19/Denmark/DCGC-13143/2020\|EPI_ISL_712613\|202 | .................... | .................... | .................... | .................... | .................... | .................... |
| X | 6217 | hCoV-19/Denmark/DCGC-13145/2020\|EPI_ISL_712614\|202 | .................... | .................... | .................... | .................... | .................... | .................... |
| X | 6218 | hCoV-19/Denmark/DCGC-13146/2020\|EPI_ISL_712615\|202 | .................... | .................... | .................... | .................... | .................... | .................... |
| X | 6219 | hCoV-19/Denmark/DCGC-13149/2020\|EPI_ISL_712616\|202 | .................... | .................... | .................... | .................... | .................... | .................... |
| X | 6220 | hCoV-19/Denmark/DCGC-13151/2020\|EPI_ISL_712617\|202 | .................... | .................... | .................... | .................... | .................... | .................... |
| X | 6221 | hCoV-19/Denmark/DCGC-13152/2020\|EPI_ISL_712618\|202 | .................... | .................... | .................... | .................... | .................... | .................... |
| X | 6222 | hCoV-19/Denmark/DCGC-13153/2020\|EPI_ISL_712619\|202 | .................... | .................... | .................... | .................... | .................... | .................... |
| X | 6223 | hCoV-19/Denmark/DCGC-13155/2020\|EPI_ISL_712620\|202 | .................... | .................... | .................... | .................... | .................... | .................... |
| X | 6224 | hCoV-19/Denmark/DCGC-13156/2020\|EPI_ISL_712621\|202 | .................... | .................... | .................... | .................... | .................... | .................... |
| X | 6225 | hCoV-19/Denmark/DCGC-13157/2020\|EPI_ISL_712622\|202 | .................... | .................... | .................... | .................... | .................... | .................... |
| X | 6226 | hCoV-19/Denmark/DCGC-13158/2020\|EPI_ISL_712623\|202 | .................... | .................... | .................... | .................... | .................... | .................... |
| X | 6227 | hCoV-19/Denmark/DCGC-13160/2020\|EPI_ISL_712624\|202 | .................... | .................... | .................... | .................... | .................... | .................... |
| X | 6228 | hCoV-19/Denmark/DCGC-13161/2020\|EPI_ISL_712625\|202 | .................... | .................... | .................... | .................... | .................... | .................... |
| X | 6229 | hCoV-19/Denmark/DCGC-13190/2020\|EPI_ISL_712640\|202 | .................... | .................... | .................... | .................... | .................... | .................... |
| X | 6230 | hCoV-19/Denmark/DCGC-13193/2020\|EPI_ISL_712641\|202 | .................... | .................... | .................... | .................... | .................... | .................... |
| X | 6231 | hCoV-19/Denmark/DCGC-13194/2020\|EPI_ISL_712642\|202 | .................... | .................... | .................... | .................... | .................... | .................... |
| X | 6232 | hCoV-19/Denmark/DCGC-13195/2020\|EPI_ISL_712643\|202 | .................... | .................... | .................... | .................... | .................... | .................... |
| X | 6233 | hCoV-19/Denmark/DCGC-13196/2020\|EPI_ISL_712644\|202 | .................... | .................... | .................... | .................... | .................... | .................... |
| X | 6234 | hCoV-19/Denmark/DCGC-13198/2020\|EPI_ISL_712645\|202 | .................... | .................... | .................... | .................... | .................... | .................... |
| X | 6235 | hCoV-19/Denmark/DCGC-13201/2020\|EPI_ISL_712648\|202 | .................... | .................... | .................... | .................... | .................... | .................... |
| X | 6236 | hCoV-19/Denmark/DCGC-13203/2020\|EPI_ISL_712649\|202 | .................... | .................... | .................... | .................... | .................... | .................... |
| X | 6237 | hCoV-19/Denmark/DCGC-13206/2020\|EPI_ISL_712650\|202 | .................... | .................... | .................... | .................... | .................... | .................... |
| X | 6238 | hCoV-19/Denmark/DCGC-13207/2020\|EPI_ISL_712651\|202 | .................... | .................... | .................... | .................... | .................... | .................... |
| X | 6239 | hCoV-19/Denmark/DCGC-13208/2020\|EPI_ISL_712652\|202 | .................... | .................... | .................... | .................... | .................... | .................... |
| X | 6240 | hCoV-19/Denmark/DCGC-13209/2020\|EPI_ISL_712653\|202 | .................... | .A.................. | .................... | .................... | .................... | ...A................ |
| X | 6241 | hCoV-19/Denmark/DCGC-13211/2020\|EPI_ISL_712654\|202 | .................... | .................... | .................... | .................... | .................... | .................... |
| X | 6242 | hCoV-19/Denmark/DCGC-13212/2020\|EPI_ISL_712655\|202 | .................... | .................... | .................... | .................... | .................... | .................... |
| X | 6243 | hCoV-19/Denmark/DCGC-13213/2020\|EPI_ISL_712656\|202 | .................... | .................... | .................... | .................... | .................... | .................... |
| X | 6244 | hCoV-19/Denmark/DCGC-13214/2020\|EPI_ISL_712657\|202 | .................... | .A.................. | .................... | .................... | .................... | .................... |
| X | 6245 | hCoV-19/Denmark/DCGC-13215/2020\|EPI_ISL_712658\|202 | .................... | .................... | .................... | .................... | .................... | .................... |
| X | 6246 | hCoV-19/Denmark/DCGC-13217/2020\|EPI_ISL_712659\|202 | .................... | .................... | .................... | .................... | .................... | .................... |
| X | 6247 | hCoV-19/Denmark/DCGC-13218/2020\|EPI_ISL_712660\|202 | .................... | .................... | .................... | .................... | .................... | .................... |
| X | 6248 | hCoV-19/Denmark/DCGC-13219/2020\|EPI_ISL_712661\|202 | .................... | .................... | .................... | .................... | .................... | .................... |
| X | 6249 | hCoV-19/Denmark/DCGC-13224/2020\|EPI_ISL_712662\|202 | .................... | .................... | .................... | .................... | .................... | .................... |
| X | 6250 | hCoV-19/Denmark/DCGC-13225/2020\|EPI_ISL_712663\|202 | .................... | .................... | .................... | .................... | .................... | .................... |
| X | 6251 | hCoV-19/Denmark/DCGC-13229/2020\|EPI_ISL_712664\|202 | .................... | .................... | .................... | .................... | .................... | .................... |
| X | 6252 | hCoV-19/Denmark/DCGC-13230/2020\|EPI_ISL_712665\|202 | .................... | .................... | .................... | .................... | .................... | .................... |
| X | 6253 | hCoV-19/Denmark/DCGC-13231/2020\|EPI_ISL_712666\|202 | .................... | .................... | .................... | .................... | .................... | .................... |
| X | 6254 | hCoV-19/Denmark/DCGC-13232/2020\|EPI_ISL_712667\|202 | .................... | .................... | .................... | .................... | .................... | .................... |
| X | 6255 | hCoV-19/Denmark/DCGC-13233/2020\|EPI_ISL_712668\|202 | .................... | .A.................. | .................... | .................... | .................... | .................... |
| X | 6256 | hCoV-19/Denmark/DCGC-13234/2020\|EPI_ISL_712669\|202 | .................... | .................... | .................... | .................... | .................... | .................... |
| X | 6257 | hCoV-19/Denmark/DCGC-13236/2020\|EPI_ISL_712670\|202 | .................... | .................... | .................... | .................... | .................... | .................... |
| X | 6258 | hCoV-19/Denmark/DCGC-13237/2020\|EPI_ISL_712671\|202 | .................... | .................... | .................... | .................... | .................... | .................... |
| X | 6259 | hCoV-19/Denmark/DCGC-13238/2020\|EPI_ISL_712672\|202 | .................... | .................... | .................... | .................... | ......T............. | .................... |
| X | 6260 | hCoV-19/Denmark/DCGC-13241/2020\|EPI_ISL_712673\|202 | .................... | .................... | .................... | .................... | .................... | .................... |

| | | | | | | | |
|---|---|---|---|---|---|---|---|
| X | 6261 hCoV-19/Denmark/DCGC-13242/2020\|EPI_ISL_712674\|202 | ···················· | ···················· | ····················· | ···················· | ···················· | ···················· |
| X | 6262 hCoV-19/Denmark/DCGC-13243/2020\|EPI_ISL_712675\|202 | ···················· | ···················· | ····················· | ···················· | ···················· | ···················· |
| X | 6263 hCoV-19/Denmark/DCGC-13244/2020\|EPI_ISL_712676\|202 | ···················· | ···················· | ····················· | ···················· | ···················· | ···················· |
| X | 6264 hCoV-19/Denmark/DCGC-13245/2020\|EPI_ISL_712677\|202 | ···················· | ···················· | ····················· | ···················· | ···················· | ···················· |
| X | 6265 hCoV-19/Denmark/DCGC-13247/2020\|EPI_ISL_712678\|202 | ···················· | ···················· | ····················· | ···················· | ···················· | ···················· |
| X | 6266 hCoV-19/Denmark/DCGC-13248/2020\|EPI_ISL_712679\|202 | ···················· | ···················· | ····················· | ···················· | ···················· | ···················· |
| X | 6267 hCoV-19/Denmark/DCGC-13249/2020\|EPI_ISL_712680\|202 | ···················· | ···················· | ····················· | ···················· | ···················· | ···················· |
| X | 6268 hCoV-19/Denmark/DCGC-13250/2020\|EPI_ISL_712681\|202 | ···················· | ···················· | ····················· | ···················· | ···················· | ···················· |
| X | 6269 hCoV-19/Denmark/DCGC-13253/2020\|EPI_ISL_712682\|202 | ···················· | ···················· | ····················· | ···················· | ···················· | ···················· |
| X | 6270 hCoV-19/Denmark/DCGC-13254/2020\|EPI_ISL_712683\|202 | ···················· | ···················· | ····················· | ···················· | ···················· | ···················· |
| X | 6271 hCoV-19/Denmark/DCGC-13255/2020\|EPI_ISL_712684\|202 | ···················· | ···················· | ····················· | ···················· | ···················· | ···················· |
| X | 6272 hCoV-19/Denmark/DCGC-13256/2020\|EPI_ISL_712685\|202 | ···················· | ···················· | ····················· | ···················· | ···················· | ···················· |
| X | 6273 hCoV-19/Denmark/DCGC-13257/2020\|EPI_ISL_712686\|202 | ···················· | ···················· | ····················· | ···················· | ···················· | ···················· |
| X | 6274 hCoV-19/Denmark/DCGC-13260/2020\|EPI_ISL_712687\|202 | ···················· | ···················· | ····················· | ···················· | ···················· | ···················· |
| X | 6275 hCoV-19/Denmark/DCGC-13261/2020\|EPI_ISL_712688\|202 | ···················· | ···················· | ····················· | ···················· | ···················· | ···················· |
| X | 6276 hCoV-19/Denmark/DCGC-13263/2020\|EPI_ISL_712689\|202 | ···················· | ···················· | ····················· | ···················· | ···················· | ···················· |
| X | 6277 hCoV-19/Denmark/DCGC-13271/2020\|EPI_ISL_712695\|202 | ···················· | ···················· | ····················· | ···················· | ···················· | ···················· |
| X | 6278 hCoV-19/Denmark/DCGC-13272/2020\|EPI_ISL_712696\|202 | ···················· | ···················· | ····················· | ···················· | ···················· | ···················· |
| X | 6279 hCoV-19/Denmark/DCGC-13274/2020\|EPI_ISL_712697\|202 | ···················· | ···················· | ····················· | ···················· | ···················· | ···················· |
| X | 6280 hCoV-19/Denmark/DCGC-13275/2020\|EPI_ISL_712698\|202 | ···················· | ···················· | ····················· | ···················· | ···················· | ···················· |
| X | 6281 hCoV-19/Denmark/DCGC-13276/2020\|EPI_ISL_712699\|202 | ···················· | ···················· | ····················· | ···················· | ···················· | ···················· |
| X | 6282 hCoV-19/Denmark/DCGC-15640/2020\|EPI_ISL_713935\|202 | ···················· | ···················· | ····················· | ···················· | ···················· | ···················· |
| X | 6283 hCoV-19/Denmark/DCGC-15643/2020\|EPI_ISL_713936\|202 | ···················· | ···················· | ····················· | ···················· | ···················· | ···················· |
| X | 6284 hCoV-19/Denmark/DCGC-15644/2020\|EPI_ISL_713937\|202 | ···················· | ···················· | ····················· | ···················· | ···················· | ···················· |
| X | 6285 hCoV-19/Denmark/DCGC-15647/2020\|EPI_ISL_713938\|202 | ···················· | ···················· | ····················· | ···················· | ···················· | ···················· |
| X | 6286 hCoV-19/Denmark/DCGC-15650/2020\|EPI_ISL_713939\|202 | ···················· | ···················· | ····················· | ···················· | ···················· | ···················· |
| X | 6287 hCoV-19/Denmark/DCGC-15651/2020\|EPI_ISL_713940\|202 | ···················· | ···················· | ····················· | ···················· | ···················· | ···················· |
| X | 6288 hCoV-19/Denmark/DCGC-15653/2020\|EPI_ISL_713941\|202 | ···················· | ···A················ | ····················· | ···················· | ···················· | ···················· |
| X | 6289 hCoV-19/Denmark/DCGC-15654/2020\|EPI_ISL_713942\|202 | ···················· | ···················· | ····················· | ···················· | ···················· | ···················· |
| X | 6290 hCoV-19/Denmark/DCGC-15657/2020\|EPI_ISL_713943\|202 | ···················· | ···················· | ····················· | ···················· | ···················· | ···················· |
| X | 6291 hCoV-19/Denmark/DCGC-15658/2020\|EPI_ISL_713944\|202 | ···················· | ···················· | ····················· | ···················· | ···················· | ···················· |
| X | 6292 hCoV-19/Denmark/DCGC-15659/2020\|EPI_ISL_713945\|202 | ···················· | ···················· | ····················· | ···················· | ···················· | ···················· |
| X | 6293 hCoV-19/Denmark/DCGC-15660/2020\|EPI_ISL_713946\|202 | ···················· | ···················· | ····················· | ···················· | ···················· | ···················· |
| X | 6294 hCoV-19/Denmark/DCGC-15662/2020\|EPI_ISL_713947\|202 | ···················· | ···················· | ····················· | ···················· | ···················· | ···················· |
| X | 6295 hCoV-19/Denmark/DCGC-15663/2020\|EPI_ISL_713948\|202 | ···················· | ···················· | ····················· | ···················· | ···················· | ···················· |
| X | 6296 hCoV-19/Denmark/DCGC-15664/2020\|EPI_ISL_713949\|202 | ···················· | ···················· | ····················· | ···················· | ···················· | ···················· |
| X | 6297 hCoV-19/Denmark/DCGC-15665/2020\|EPI_ISL_713950\|202 | ···················· | ···················· | ····················· | ···················· | ···················· | ···················· |
| X | 6298 hCoV-19/Denmark/DCGC-15666/2020\|EPI_ISL_713951\|202 | ···················· | ···················· | ····················· | ···················· | ···················· | ···················· |
| X | 6299 hCoV-19/Denmark/DCGC-15667/2020\|EPI_ISL_713952\|202 | ···················· | ···················· | ····················· | ···················· | ···················· | ···················· |
| X | 6300 hCoV-19/Denmark/DCGC-15668/2020\|EPI_ISL_713953\|202 | ···················· | ···················· | ····················· | ···················· | ···················· | ···················· |
| X | 6301 hCoV-19/Denmark/DCGC-15669/2020\|EPI_ISL_713954\|202 | ···················· | ···················· | ····················· | ···················· | ···················· | ···················· |
| X | 6302 hCoV-19/Denmark/DCGC-15673/2020\|EPI_ISL_713955\|202 | ···················· | ···················· | ····················· | ···················· | ···················· | ···················· |
| X | 6303 hCoV-19/Denmark/DCGC-15674/2020\|EPI_ISL_713956\|202 | ···················· | ···················· | ····················· | ···················· | ···················· | ···················· |
| X | 6304 hCoV-19/Denmark/DCGC-15700/2020\|EPI_ISL_713970\|202 | ···················· | ···················· | ····················· | ···················· | ···················· | ···················· |
| X | 6305 hCoV-19/Denmark/DCGC-15701/2020\|EPI_ISL_713971\|202 | ···················· | ···················· | ····················· | ···················· | ···················· | ···················· |
| X | 6306 hCoV-19/Denmark/DCGC-15703/2020\|EPI_ISL_713972\|202 | ···················· | ···················· | ····················· | ···················· | ···················· | ···················· |
| X | 6307 hCoV-19/Denmark/DCGC-15705/2020\|EPI_ISL_713973\|202 | ···················· | ···················· | ····················· | ···················· | ···················· | ···················· |
| X | 6308 hCoV-19/Denmark/DCGC-15707/2020\|EPI_ISL_713974\|202 | ···················· | ···················· | ····················· | ···················· | ···················· | ···················· |
| X | 6309 hCoV-19/Denmark/DCGC-15710/2020\|EPI_ISL_713975\|202 | ···················· | ···················· | ····················· | ···················· | ···················· | ···················· |
| X | 6310 hCoV-19/Denmark/DCGC-15717/2020\|EPI_ISL_713979\|202 | ···················· | ···················· | ····················· | ···················· | ···················· | ···················· |
| X | 6311 hCoV-19/Denmark/DCGC-15718/2020\|EPI_ISL_713980\|202 | ···················· | ·A·················· | ····················· | ···················· | ···················· | ··A················· |
| X | 6312 hCoV-19/Denmark/DCGC-15720/2020\|EPI_ISL_713981\|202 | ···················· | ···················· | ····················· | ···················· | ···················· | ···················· |
| X | 6313 hCoV-19/Denmark/DCGC-15722/2020\|EPI_ISL_713982\|202 | ···················· | ···················· | ····················· | ···················· | ···················· | ···················· |
| X | 6314 hCoV-19/Denmark/DCGC-15725/2020\|EPI_ISL_713983\|202 | ···················· | ···················· | ····················· | ···················· | ···················· | ···················· |
| X | 6315 hCoV-19/Denmark/DCGC-15726/2020\|EPI_ISL_713984\|202 | ···················· | ···················· | ····················· | ···················· | ···················· | ···················· |
| X | 6316 hCoV-19/Denmark/DCGC-15727/2020\|EPI_ISL_713985\|202 | ···················· | ···················· | ····················· | ···················· | ···················· | ···················· |
| X | 6317 hCoV-19/Denmark/DCGC-15729/2020\|EPI_ISL_713986\|202 | ···················· | ···················· | ····················· | ···················· | ···················· | ···················· |
| X | 6318 hCoV-19/Denmark/DCGC-15732/2020\|EPI_ISL_713987\|202 | ···················· | ···················· | ····················· | ···················· | ···················· | ···················· |
| X | 6319 hCoV-19/Denmark/DCGC-15734/2020\|EPI_ISL_713988\|202 | ···················· | ···················· | ····················· | ···················· | ···················· | ···················· |
| X | 6320 hCoV-19/Denmark/DCGC-15736/2020\|EPI_ISL_713989\|202 | ···················· | ·A·················· | ····················· | ···················· | ···················· | ···················· |
| X | 6321 hCoV-19/Denmark/DCGC-15738/2020\|EPI_ISL_713990\|202 | ···················· | ···················· | ····················· | ···················· | ···················· | ···················· |
| X | 6322 hCoV-19/Denmark/DCGC-15739/2020\|EPI_ISL_713991\|202 | ···················· | ···················· | ····················· | ···················· | ···················· | ···················· |
| X | 6323 hCoV-19/Denmark/DCGC-15740/2020\|EPI_ISL_713992\|202 | ···················· | ···················· | ····················· | ···················· | ···················· | ···················· |
| X | 6324 hCoV-19/Denmark/DCGC-15741/2020\|EPI_ISL_713993\|202 | ···················· | ···················· | ····················· | ···················· | ···················· | ···················· |
| X | 6325 hCoV-19/Denmark/DCGC-15743/2020\|EPI_ISL_713994\|202 | ···················· | ···················· | ····················· | ···················· | ···················· | ···················· |
| X | 6326 hCoV-19/Denmark/DCGC-15744/2020\|EPI_ISL_713995\|202 | ···················· | ···················· | ····················· | ···················· | ···················· | ···················· |
| X | 6327 hCoV-19/Denmark/DCGC-15745/2020\|EPI_ISL_713996\|202 | ···················· | ···················· | ····················· | ···················· | ···················· | ···················· |
| X | 6328 hCoV-19/Denmark/DCGC-15749/2020\|EPI_ISL_713997\|202 | ···················· | ···················· | ····················· | ···················· | ···················· | ···················· |
| X | 6329 hCoV-19/Denmark/DCGC-15751/2020\|EPI_ISL_713998\|202 | ···················· | ···················· | ····················· | ···················· | ···················· | ···················· |
| X | 6330 hCoV-19/Denmark/DCGC-15752/2020\|EPI_ISL_713999\|202 | ···················· | ···················· | ····················· | ···················· | ···················· | ···················· |
| X | 6331 hCoV-19/Netherlands/OV-RIVM-20125/2020\|EPI_ISL_723 | ···················· | ···················· | ····················· | ···················· | ···················· | ···················· |
| X | 6332 hCoV-19/Netherlands/NH-RIVM-20128/2020\|EPI_ISL_723 | ···················· | ···················· | ····················· | ···················· | ···················· | ···················· |
| X | 6333 hCoV-19/Netherlands/NH-RIVM-20129/2020\|EPI_ISL_723 | ···················· | ···················· | ····················· | ···················· | ···················· | ···················· |
| X | 6334 hCoV-19/Netherlands/NH-RIVM-20130/2020\|EPI_ISL_723 | ···················· | ···················· | ····················· | ···················· | ···················· | ···················· |
| X | 6335 hCoV-19/Netherlands/NH-RIVM-20133/2020\|EPI_ISL_723 | ···················· | ···················· | ····················· | ···················· | ···················· | ···················· |
| X | 6336 hCoV-19/Netherlands/NH-RIVM-20134/2020\|EPI_ISL_723 | ···················· | ···················· | ····················· | ···················· | ···················· | ···················· |
| X | 6337 hCoV-19/Netherlands/UT-RIVM-20135/2020\|EPI_ISL_723 | ···················· | ···················· | ····················· | ···················· | ·····T·············· | ···················· |
| X | 6338 hCoV-19/Netherlands/UT-RIVM-20136/2020\|EPI_ISL_723 | ···················· | ···················· | ····················· | ···················· | ···················· | ···················· |
| X | 6339 hCoV-19/Netherlands/UT-RIVM-20137/2020\|EPI_ISL_723 | ···················· | ···················· | ····················· | ···················· | ·····T·············· | ···················· |
| X | 6340 hCoV-19/Netherlands/UT-RIVM-20138/2020\|EPI_ISL_723 | ···················· | ···················· | ····················· | ···················· | ···················· | ···················· |

| | | | | | | | | |
|---|---|---|---|---|---|---|---|---|
| X | 6341 | hCoV-19/Netherlands/UT-RIVM-20140/2020\|EPI_ISL_723 | ···················· | ···················· | ····················· | ···················· | ···················· | ···················· |
| X | 6342 | hCoV-19/Netherlands/UT-RIVM-20141/2020\|EPI_ISL_723 | ···················· | ···················· | ····················· | ···················· | ···················· | ···················· |
| X | 6343 | hCoV-19/Netherlands/UT-RIVM-20142/2020\|EPI_ISL_723 | ···················· | ···················· | ····················· | ···················· | ···················· | ···················· |
| X | 6344 | hCoV-19/Netherlands/OV-RIVM-20143/2020\|EPI_ISL_723 | ···················· | ···················· | ····················· | ···················· | ···················· | ···················· |
| X | 6345 | hCoV-19/Netherlands/OV-RIVM-20144/2020\|EPI_ISL_723 | ···················· | ···················· | ····················· | ···················· | ···················· | ···················· |
| X | 6346 | hCoV-19/Netherlands/OV-RIVM-20145/2020\|EPI_ISL_723 | ···················· | ···················· | ····················· | ···················· | ···················· | ···················· |
| X | 6347 | hCoV-19/Netherlands/FR-RIVM-20146/2020\|EPI_ISL_723 | ···················· | ···················· | ····················· | ···················· | ···················· | ···················· |
| X | 6348 | hCoV-19/Netherlands/FR-RIVM-20147/2020\|EPI_ISL_723 | ···················· | ···················· | ····················· | ···················· | ···················· | ···················· |
| X | 6349 | hCoV-19/Netherlands/GE-RIVM-20148/2020\|EPI_ISL_723 | ···················· | ···················· | ····················· | ···················· | ······T············· | ···················· |
| X | 6350 | hCoV-19/Netherlands/UT-RIVM-20149/2020\|EPI_ISL_723 | ···················· | ···················· | ····················· | ···················· | ···················· | ···················· |
| X | 6351 | hCoV-19/Netherlands/UT-RIVM-20150/2020\|EPI_ISL_723 | ···················· | ···················· | ····················· | ···················· | ···················· | ···················· |
| X | 6352 | hCoV-19/Netherlands/OV-RIVM-20151/2020\|EPI_ISL_723 | ···················· | ······A············· | ····················· | ···················· | ···················· | ···················· |
| X | 6353 | hCoV-19/Netherlands/OV-RIVM-20152/2020\|EPI_ISL_723 | ···················· | ···················· | ····················· | ···················· | ···················· | ···················· |
| X | 6354 | hCoV-19/Netherlands/FR-RIVM-20153/2020\|EPI_ISL_723 | ···················· | ···················· | ····················· | ···················· | ···················· | ···················· |
| X | 6355 | hCoV-19/Netherlands/OV-RIVM-20157/2020\|EPI_ISL_723 | ···················· | ···················· | ····················· | ···················· | ···················· | ···················· |
| X | 6356 | hCoV-19/Netherlands/OV-RIVM-20158/2020\|EPI_ISL_723 | ···················· | ···················· | ····················· | ···················· | ···················· | ···················· |
| X | 6357 | hCoV-19/Netherlands/GE-RIVM-20159/2020\|EPI_ISL_723 | ···················· | ···················· | ····················· | ···················· | ···················· | ···················· |
| X | 6358 | hCoV-19/Netherlands/NH-RIVM-20160/2020\|EPI_ISL_723 | ···················· | ···················· | ····················· | ···················· | ···················· | ···················· |
| X | 6359 | hCoV-19/Netherlands/UT-RIVM-20161/2020\|EPI_ISL_723 | ···················· | ···················· | ····················· | ···················· | ···················· | ···················· |
| X | 6360 | hCoV-19/Netherlands/UT-RIVM-20162/2020\|EPI_ISL_723 | ···················· | ···················· | ····················· | ···················· | ···················· | ···················· |
| X | 6361 | hCoV-19/Netherlands/UT-RIVM-20163/2020\|EPI_ISL_723 | ···················· | ···················· | ····················· | ···················· | ···················· | ···················· |
| X | 6362 | hCoV-19/Netherlands/UT-RIVM-20164/2020\|EPI_ISL_723 | ···················· | ···················· | ····················· | ···················· | ···················· | ···················· |
| X | 6363 | hCoV-19/Netherlands/UT-RIVM-20166/2020\|EPI_ISL_723 | ···················· | ···················· | ····················· | ···················· | ···················· | ···················· |
| X | 6364 | hCoV-19/Netherlands/ZH-RIVM-20167/2020\|EPI_ISL_723 | ···················· | ···················· | ····················· | ···················· | ···················· | ···················· |
| X | 6365 | hCoV-19/Netherlands/OV-RIVM-20168/2020\|EPI_ISL_723 | ···················· | ···················· | ····················· | ···················· | ···················· | ···················· |
| X | 6366 | hCoV-19/Netherlands/UT-RIVM-20174/2020\|EPI_ISL_723 | ···················· | ···················· | ····················· | ···················· | ···················· | ···················· |
| X | 6367 | hCoV-19/Netherlands/UT-RIVM-20175/2020\|EPI_ISL_723 | ···················· | ···················· | ····················· | ···················· | ···················· | ···················· |
| X | 6368 | hCoV-19/Netherlands/UT-RIVM-20176/2020\|EPI_ISL_723 | ···················· | ···················· | ····················· | ···················· | ···················· | ···················· |
| X | 6369 | hCoV-19/Netherlands/UT-RIVM-20177/2020\|EPI_ISL_723 | ···················· | ···················· | ····················· | ···················· | ···················· | ···················· |
| X | 6370 | hCoV-19/Netherlands/UT-RIVM-20178/2020\|EPI_ISL_723 | ···················· | ···················· | ····················· | ···················· | ···················· | ···················· |
| X | 6371 | hCoV-19/Netherlands/NH-RIVM-20185/2020\|EPI_ISL_723 | ···················· | ···················· | ····················· | ···················· | ···················· | ···················· |
| X | 6372 | hCoV-19/Netherlands/NH-RIVM-20186/2020\|EPI_ISL_723 | ···················· | ···················· | ····················· | ···················· | ···················· | ···················· |
| X | 6373 | hCoV-19/Netherlands/UT-RIVM-20188/2020\|EPI_ISL_723 | ···················· | ···················· | ····················· | ···················· | ···················· | ···················· |
| X | 6374 | hCoV-19/Netherlands/UT-RIVM-20189/2020\|EPI_ISL_723 | ···················· | ···················· | ····················· | ···················· | ···················· | ···················· |
| X | 6375 | hCoV-19/Netherlands/UT-RIVM-20190/2020\|EPI_ISL_723 | ···················· | ···················· | ····················· | ···················· | ···················· | ···················· |
| X | 6376 | hCoV-19/Netherlands/UT-RIVM-20191/2020\|EPI_ISL_723 | ···················· | ···················· | ····················· | ···················· | ···················· | ···················· |
| X | 6377 | hCoV-19/Netherlands/UT-RIVM-20192/2020\|EPI_ISL_723 | ···················· | ···················· | ····················· | ···················· | ···················· | ···················· |
| X | 6378 | hCoV-19/Netherlands/UT-RIVM-20193/2020\|EPI_ISL_723 | ···················· | ···················· | ····················· | ···················· | ···················· | ···················· |
| X | 6379 | hCoV-19/Netherlands/FR-RIVM-20194/2020\|EPI_ISL_723 | ···················· | ···················· | ····················· | ···················· | ···················· | ···················· |
| X | 6380 | hCoV-19/Netherlands/FR-RIVM-20195/2020\|EPI_ISL_723 | ···················· | ···················· | ····················· | ···················· | ···················· | ···················· |
| X | 6381 | hCoV-19/Netherlands/GE-RIVM-20207/2020\|EPI_ISL_723 | ···················· | ···················· | ····················· | ···················· | ···················· | ···················· |
| X | 6382 | hCoV-19/Netherlands/GE-RIVM-20208/2020\|EPI_ISL_723 | ···················· | ···················· | ····················· | ···················· | ···················· | ···················· |
| X | 6383 | hCoV-19/Netherlands/GE-RIVM-20209/2020\|EPI_ISL_723 | ···················· | ···················· | ····················· | ···················· | ···················· | ···················· |
| X | 6384 | hCoV-19/Netherlands/NB-RIVM-20210/2020\|EPI_ISL_723 | ···················· | ···················· | ····················· | ···················· | ···················· | ···················· |
| X | 6385 | hCoV-19/Netherlands/LI-RIVM-20211/2020\|EPI_ISL_723 | ···················· | ···················· | ····················· | ···················· | ···················· | ···················· |
| X | 6386 | hCoV-19/Netherlands/FR-RIVM-20212/2020\|EPI_ISL_723 | ···················· | ···················· | ····················· | ···················· | ···················· | ···················· |
| X | 6387 | hCoV-19/Netherlands/LI-RIVM-20213/2020\|EPI_ISL_723 | ···················· | ···················· | ····················· | ···················· | ···················· | ···················· |
| X | 6388 | hCoV-19/Netherlands/GE-RIVM-20214/2020\|EPI_ISL_723 | ···················· | ···················· | ····················· | ···················· | ···················· | ···················· |
| X | 6389 | hCoV-19/Netherlands/LI-RIVM-20215/2020\|EPI_ISL_723 | ···················· | ···················· | ····················· | ···················· | ···················· | ···················· |
| X | 6390 | hCoV-19/Netherlands/GE-RIVM-20216/2020\|EPI_ISL_723 | ···················· | ···················· | ····················· | ···················· | ···················· | ···················· |
| X | 6391 | hCoV-19/Netherlands/NH-RIVM-20217/2020\|EPI_ISL_723 | ···················· | ···················· | ····················· | ···················· | ···················· | ···················· |
| X | 6392 | hCoV-19/Netherlands/NH-RIVM-20218/2020\|EPI_ISL_723 | ···················· | ···················· | ····················· | ···················· | ···················· | ···················· |
| X | 6393 | hCoV-19/Netherlands/NH-RIVM-20219/2020\|EPI_ISL_723 | ···················· | ···················· | ····················· | ···················· | ···················· | ···················· |
| X | 6394 | hCoV-19/Netherlands/UT-RIVM-20220/2020\|EPI_ISL_723 | ···················· | ···················· | ····················· | ···················· | ···················· | ···················· |
| X | 6395 | hCoV-19/Netherlands/NH-RIVM-20222/2020\|EPI_ISL_723 | ···················· | ···················· | ····················· | ···················· | ···················· | ···················· |
| X | 6396 | hCoV-19/Netherlands/NH-RIVM-20223/2020\|EPI_ISL_723 | ···················· | ···················· | ····················· | ···················· | ···················· | ···················· |
| X | 6397 | hCoV-19/Netherlands/NH-RIVM-20224/2020\|EPI_ISL_723 | ···················· | ···················· | ····················· | ···················· | ···················· | ···················· |
| X | 6398 | hCoV-19/Netherlands/NH-RIVM-20225/2020\|EPI_ISL_723 | ···················· | ···················· | ····················· | ···················· | ···················· | ···················· |
| X | 6399 | hCoV-19/Netherlands/NH-RIVM-20226/2020\|EPI_ISL_723 | ···················· | ·············A······ | ····················· | ···················· | ···················· | ···················· |
| X | 6400 | hCoV-19/Netherlands/NH-RIVM-20229/2020\|EPI_ISL_723 | ···················· | ···················· | ····················· | ···················· | ···················· | ···················· |
| X | 6401 | hCoV-19/Denmark/DCGC-14561/2020\|EPI_ISL_711360\|202 | ···················· | ···················· | ····················· | ···················· | ···················· | ···················· |
| X | 6402 | hCoV-19/Denmark/DCGC-14599/2020\|EPI_ISL_711361\|202 | ···················· | ···················· | ····················· | ···················· | ···················· | ···················· |
| X | 6403 | hCoV-19/Denmark/DCGC-14614/2020\|EPI_ISL_711362\|202 | ···················· | ···················· | ····················· | ···················· | ···················· | ···················· |
| X | 6404 | hCoV-19/Denmark/DCGC-14624/2020\|EPI_ISL_711363\|202 | ···················· | ···················· | ····················· | ············A······· | ···················· | ···················· |
| X | 6405 | hCoV-19/Denmark/DCGC-14764/2020\|EPI_ISL_711370\|202 | ···················· | ···················· | ····················· | ···················· | ···················· | ···················· |
| X | 6406 | hCoV-19/Denmark/DCGC-14783/2020\|EPI_ISL_711371\|202 | ···················· | ···················· | ····················· | ···················· | ···················· | ···················· |
| X | 6407 | hCoV-19/Denmark/DCGC-14788/2020\|EPI_ISL_711372\|202 | ···················· | ···················· | ····················· | ···················· | ···················· | ···················· |
| X | 6408 | hCoV-19/Denmark/DCGC-14798/2020\|EPI_ISL_711373\|202 | ···················· | ···················· | ····················· | ···················· | ···················· | ···················· |
| X | 6409 | hCoV-19/Denmark/DCGC-14809/2020\|EPI_ISL_711374\|202 | ···················· | ···················· | ····················· | ···················· | ···················· | ···················· |
| X | 6410 | hCoV-19/Denmark/DCGC-14811/2020\|EPI_ISL_711375\|202 | ···················· | ···················· | ····················· | ···················· | ···················· | ···················· |
| X | 6411 | hCoV-19/Denmark/DCGC-14842/2020\|EPI_ISL_711376\|202 | ···················· | ···················· | ····················· | ···················· | ···················· | ···················· |
| X | 6412 | hCoV-19/Denmark/DCGC-14846/2020\|EPI_ISL_711377\|202 | ···················· | ···················· | ····················· | ···················· | ···················· | ···················· |
| X | 6413 | hCoV-19/Denmark/DCGC-14870/2020\|EPI_ISL_711378\|202 | ···················· | ···················· | ····················· | ···················· | ···C················ | ···················· |
| X | 6414 | hCoV-19/Denmark/DCGC-14878/2020\|EPI_ISL_711379\|202 | ···················· | ···················· | ····················· | ···················· | ···················· | ···················· |
| X | 6415 | hCoV-19/Denmark/DCGC-13911/2020\|EPI_ISL_711445\|202 | ···················· | ···················· | ····················· | ···················· | ···················· | ···················· |
| X | 6416 | hCoV-19/Denmark/DCGC-13912/2020\|EPI_ISL_711446\|202 | ···················· | ···················· | ····················· | ···················· | ···················· | ···················· |
| X | 6417 | hCoV-19/Denmark/DCGC-13914/2020\|EPI_ISL_711447\|202 | ···················· | ···················· | ····················· | ···················· | ···················· | ···················· |
| X | 6418 | hCoV-19/Denmark/DCGC-13915/2020\|EPI_ISL_711448\|202 | ···················· | ···················· | ····················· | ···················· | ···················· | ···················· |
| X | 6419 | hCoV-19/Denmark/DCGC-13930/2020\|EPI_ISL_711460\|202 | ···················· | ···················· | ····················· | ···················· | ···················· | ···················· |
| X | 6420 | hCoV-19/Denmark/DCGC-13931/2020\|EPI_ISL_711461\|202 | ···················· | ···················· | ····················· | ···················· | ···················· | ···················· |

| | | | | | | | |
|---|---|---|---|---|---|---|---|
| X | 6421 hCoV-19/Denmark/DCGC-13932/2020\|EPI_ISL_711462\|202 | ···················· | ·A·················· | ···················· | ···················· | ···················· | ··A················· |
| X | 6422 hCoV-19/Denmark/DCGC-13933/2020\|EPI_ISL_711463\|202 | ···················· | ···················· | ···················· | ···················· | ···················· | ···················· |
| X | 6423 hCoV-19/Denmark/DCGC-13935/2020\|EPI_ISL_711464\|202 | ···················· | ···················· | ···················· | ···················· | ···················· | ···················· |
| X | 6424 hCoV-19/Denmark/DCGC-13940/2020\|EPI_ISL_711465\|202 | ···················· | ···················· | ···················· | ···················· | ···················· | ···················· |
| X | 6425 hCoV-19/Denmark/DCGC-13944/2020\|EPI_ISL_711466\|202 | ···················· | ···················· | ···················· | ···················· | ···················· | ···················· |
| X | 6426 hCoV-19/Denmark/DCGC-13945/2020\|EPI_ISL_711467\|202 | ···················· | ···················· | ···················· | ···················· | ···················· | ···················· |
| X | 6427 hCoV-19/Denmark/DCGC-13947/2020\|EPI_ISL_711468\|202 | ···················· | ···················· | ···················· | ···················· | ···················· | ···················· |
| X | 6428 hCoV-19/Denmark/DCGC-13948/2020\|EPI_ISL_711469\|202 | ···················· | ···················· | ···················· | ···················· | ···················· | ···················· |
| X | 6429 hCoV-19/Denmark/DCGC-13949/2020\|EPI_ISL_711470\|202 | ···················· | ···················· | ···················· | ···················· | ···················· | ···················· |
| X | 6430 hCoV-19/Denmark/DCGC-13950/2020\|EPI_ISL_711471\|202 | ···················· | ···················· | ···················· | ···················· | ···················· | ···················· |
| X | 6431 hCoV-19/Denmark/DCGC-13952/2020\|EPI_ISL_711472\|202 | ···················· | ···················· | ···················· | ···················· | ···················· | ···················· |
| X | 6432 hCoV-19/Denmark/DCGC-13953/2020\|EPI_ISL_711473\|202 | ···················· | ···················· | ···················· | ···················· | ···················· | ···················· |
| X | 6433 hCoV-19/Denmark/DCGC-13954/2020\|EPI_ISL_711474\|202 | ···················· | ···················· | ···················· | ···················· | ···················· | ···················· |
| X | 6434 hCoV-19/Denmark/DCGC-13958/2020\|EPI_ISL_711475\|202 | ···················· | ···················· | ···················· | ···················· | ···················· | ···················· |
| X | 6435 hCoV-19/Denmark/DCGC-13959/2020\|EPI_ISL_711476\|202 | ···················· | ···················· | ···················· | ···················· | ···················· | ···················· |
| X | 6436 hCoV-19/Denmark/DCGC-13960/2020\|EPI_ISL_711477\|202 | ···················· | ···················· | ···················· | ···················· | ···················· | ···················· |
| X | 6437 hCoV-19/Denmark/DCGC-13961/2020\|EPI_ISL_711478\|202 | ···················· | ···················· | ···················· | ···················· | ···················· | ···················· |
| X | 6438 hCoV-19/Denmark/DCGC-13962/2020\|EPI_ISL_711479\|202 | ···················· | ···················· | ···················· | ···················· | ···················· | ···················· |
| X | 6439 hCoV-19/Denmark/DCGC-13265/2020\|EPI_ISL_712690\|202 | ···················· | ···················· | ···················· | ···················· | ···················· | ···················· |
| X | 6440 hCoV-19/Denmark/DCGC-13266/2020\|EPI_ISL_712691\|202 | ···················· | ···················· | ···················· | ···················· | ···················· | ···················· |
| X | 6441 hCoV-19/Denmark/DCGC-13267/2020\|EPI_ISL_712692\|202 | ···················· | ···················· | ···················· | ···················· | ···················· | ···················· |
| X | 6442 hCoV-19/Denmark/DCGC-13268/2020\|EPI_ISL_712693\|202 | ···················· | ···················· | ···················· | ···················· | ···················· | ···················· |
| X | 6443 hCoV-19/Denmark/DCGC-13270/2020\|EPI_ISL_712694\|202 | ···················· | ···················· | ···················· | ···················· | ···················· | ···················· |
| X | 6444 hCoV-19/Denmark/DCGC-13278/2020\|EPI_ISL_712700\|202 | ···················· | ···················· | ···················· | ···················· | ···················· | ···················· |
| X | 6445 hCoV-19/Denmark/DCGC-13280/2020\|EPI_ISL_712701\|202 | ···················· | ···················· | ···················· | ···················· | ···················· | ···················· |
| X | 6446 hCoV-19/Denmark/DCGC-13282/2020\|EPI_ISL_712702\|202 | ···················· | ···················· | ···················· | ···················· | ···················· | ···················· |
| X | 6447 hCoV-19/Denmark/DCGC-13283/2020\|EPI_ISL_712703\|202 | ···················· | ···················· | ···················· | ···················· | ···················· | ···················· |
| X | 6448 hCoV-19/Denmark/DCGC-13284/2020\|EPI_ISL_712704\|202 | ···················· | ···················· | ···················· | ···················· | ···················· | ···················· |
| X | 6449 hCoV-19/Denmark/DCGC-13285/2020\|EPI_ISL_712705\|202 | ···················· | ···················· | ···················· | ···················· | ···················· | ···················· |
| X | 6450 hCoV-19/Denmark/DCGC-13286/2020\|EPI_ISL_712706\|202 | ···················· | ···················· | ···················· | ···················· | ···················· | ···················· |
| X | 6451 hCoV-19/Denmark/DCGC-13287/2020\|EPI_ISL_712707\|202 | ···················· | ···················· | ···················· | ···················· | ···················· | ···················· |
| X | 6452 hCoV-19/Denmark/DCGC-13288/2020\|EPI_ISL_712708\|202 | ···················· | ···················· | ···················· | ···················· | ···················· | ···················· |
| X | 6453 hCoV-19/Denmark/DCGC-13289/2020\|EPI_ISL_712709\|202 | ···················· | ···················· | ···················· | ···················· | ···················· | ···················· |
| X | 6454 hCoV-19/Denmark/DCGC-13290/2020\|EPI_ISL_712710\|202 | ···················· | ···················· | ···················· | ···················· | ···················· | ···················· |
| X | 6455 hCoV-19/Denmark/DCGC-13291/2020\|EPI_ISL_712711\|202 | ···················· | ···················· | ···················· | ···················· | ···················· | ···················· |
| X | 6456 hCoV-19/Denmark/DCGC-13292/2020\|EPI_ISL_712712\|202 | ···················· | ···················· | ···················· | ···················· | ···················· | ···················· |
| X | 6457 hCoV-19/Denmark/DCGC-13293/2020\|EPI_ISL_712713\|202 | ···················· | ···················· | ···················· | ···················· | ···················· | ···················· |
| X | 6458 hCoV-19/Denmark/DCGC-13294/2020\|EPI_ISL_712714\|202 | ···················· | ···················· | ···················· | ···················· | ···················· | ···················· |
| X | 6459 hCoV-19/Denmark/DCGC-13295/2020\|EPI_ISL_712715\|202 | ···················· | ···················· | ···················· | ···················· | ···················· | ···················· |
| X | 6460 hCoV-19/Denmark/DCGC-13296/2020\|EPI_ISL_712716\|202 | ···················· | ···················· | ···················· | ···················· | ···················· | ···················· |
| X | 6461 hCoV-19/Denmark/DCGC-13297/2020\|EPI_ISL_712717\|202 | ···················· | ···················· | ···················· | ···················· | ···················· | ···················· |
| X | 6462 hCoV-19/Denmark/DCGC-13299/2020\|EPI_ISL_712718\|202 | ···················· | ···················· | ···················· | ···················· | ···················· | ···················· |
| X | 6463 hCoV-19/Denmark/DCGC-13302/2020\|EPI_ISL_712719\|202 | ···················· | ···················· | ···················· | ···················· | ···················· | ···················· |
| X | 6464 hCoV-19/Denmark/DCGC-13305/2020\|EPI_ISL_712720\|202 | ···················· | ···················· | ···················· | ···················· | ···················· | ···················· |
| X | 6465 hCoV-19/Denmark/DCGC-13307/2020\|EPI_ISL_712721\|202 | ···················· | ···················· | ···················· | ···················· | ···················· | ···················· |
| X | 6466 hCoV-19/Denmark/DCGC-13312/2020\|EPI_ISL_712722\|202 | ···················· | ···················· | ···················· | ···················· | ···················· | ···················· |
| X | 6467 hCoV-19/Denmark/DCGC-13313/2020\|EPI_ISL_712723\|202 | ···················· | ···················· | ···················· | ···················· | ···················· | ···················· |
| X | 6468 hCoV-19/Denmark/DCGC-13315/2020\|EPI_ISL_712724\|202 | ···················· | ···················· | ···················· | ···················· | ···················· | ···················· |
| X | 6469 hCoV-19/Denmark/DCGC-13317/2020\|EPI_ISL_712725\|202 | ···················· | ···················· | ···················· | ···················· | ···················· | ···················· |
| X | 6470 hCoV-19/Denmark/DCGC-13318/2020\|EPI_ISL_712726\|202 | ···················· | ···················· | ···················· | ···················· | ···················· | ···················· |
| X | 6471 hCoV-19/Denmark/DCGC-13320/2020\|EPI_ISL_712727\|202 | ···················· | ···················· | ···················· | ···················· | ···················· | ···················· |
| X | 6472 hCoV-19/Denmark/DCGC-13323/2020\|EPI_ISL_712728\|202 | ···················· | ···················· | ···················· | ···················· | ···················· | ···················· |
| X | 6473 hCoV-19/Denmark/DCGC-13324/2020\|EPI_ISL_712729\|202 | ···················· | ···················· | ···················· | ···················· | ···················· | ···················· |
| X | 6474 hCoV-19/Denmark/DCGC-13326/2020\|EPI_ISL_712730\|202 | ···················· | ···················· | ···················· | ···················· | ···················· | ···················· |
| X | 6475 hCoV-19/Denmark/DCGC-13327/2020\|EPI_ISL_712731\|202 | ···················· | ···················· | ···················· | ···················· | ···················· | ···················· |
| X | 6476 hCoV-19/Denmark/DCGC-13328/2020\|EPI_ISL_712732\|202 | ···················· | ···················· | ···················· | ···················· | ···················· | ···················· |
| X | 6477 hCoV-19/Denmark/DCGC-13331/2020\|EPI_ISL_712733\|202 | ···················· | ···················· | ···················· | ···················· | ···················· | ···················· |
| X | 6478 hCoV-19/Denmark/DCGC-13332/2020\|EPI_ISL_712734\|202 | ···················· | ···················· | ···················· | ···················· | ···················· | ···················· |
| X | 6479 hCoV-19/Denmark/DCGC-13333/2020\|EPI_ISL_712735\|202 | ···················· | ···················· | ···················· | ···················· | ···················· | ···················· |
| X | 6480 hCoV-19/Denmark/DCGC-13335/2020\|EPI_ISL_712736\|202 | ···················· | ···················· | ···················· | ···················· | ···················· | ···················· |
| X | 6481 hCoV-19/Denmark/DCGC-13336/2020\|EPI_ISL_712737\|202 | ···················· | ···················· | ···················· | ···················· | ···················· | ···················· |
| X | 6482 hCoV-19/Denmark/DCGC-13337/2020\|EPI_ISL_712738\|202 | ···················· | ···················· | ···················· | ···················· | ···················· | ···················· |
| X | 6483 hCoV-19/Denmark/DCGC-13338/2020\|EPI_ISL_712739\|202 | ···················· | ···················· | ···················· | ···················· | ···················· | ···················· |
| X | 6484 hCoV-19/Denmark/DCGC-13340/2020\|EPI_ISL_712740\|202 | ···················· | ···················· | ···················· | ···················· | ···················· | ···················· |
| X | 6485 hCoV-19/Denmark/DCGC-13341/2020\|EPI_ISL_712741\|202 | ···················· | ···················· | ···················· | ···················· | ···················· | ···················· |
| X | 6486 hCoV-19/Denmark/DCGC-13342/2020\|EPI_ISL_712742\|202 | ···················· | ···················· | ···················· | ···················· | ···················· | ···················· |
| X | 6487 hCoV-19/Denmark/DCGC-13349/2020\|EPI_ISL_712743\|202 | ···················· | ···················· | ···················· | ···················· | ···················· | ···················· |
| X | 6488 hCoV-19/Denmark/DCGC-13350/2020\|EPI_ISL_712744\|202 | ···················· | ···················· | ···················· | ···················· | ···················· | ···················· |
| X | 6489 hCoV-19/Denmark/DCGC-13353/2020\|EPI_ISL_712745\|202 | ···················· | ···················· | ···················· | ···················· | ···················· | ···················· |
| X | 6490 hCoV-19/Denmark/DCGC-13355/2020\|EPI_ISL_712746\|202 | ···················· | ···················· | ···················· | ···················· | ···················· | ···················· |
| X | 6491 hCoV-19/Denmark/DCGC-13358/2020\|EPI_ISL_712747\|202 | ···················· | ···················· | ···················· | ···················· | ···················· | ···················· |
| X | 6492 hCoV-19/Denmark/DCGC-13359/2020\|EPI_ISL_712748\|202 | ···················· | ···················· | ···················· | ···················· | ···················· | ···················· |
| X | 6493 hCoV-19/Denmark/DCGC-13361/2020\|EPI_ISL_712749\|202 | ···················· | ···················· | ···················· | ···················· | ···················· | ···················· |
| X | 6494 hCoV-19/Denmark/DCGC-13363/2020\|EPI_ISL_712750\|202 | ···················· | ···················· | ···················· | ···················· | ···················· | ···················· |
| X | 6495 hCoV-19/Denmark/DCGC-13364/2020\|EPI_ISL_712751\|202 | ···················· | ···················· | ···················· | ···················· | ···················· | ···················· |
| X | 6496 hCoV-19/Denmark/DCGC-13365/2020\|EPI_ISL_712752\|202 | ···················· | ···················· | ···················· | ···················· | ···················· | ···················· |
| X | 6497 hCoV-19/Denmark/DCGC-13367/2020\|EPI_ISL_712753\|202 | ···················· | ···················· | ···················· | ···················· | ···················· | ···················· |
| X | 6498 hCoV-19/Denmark/DCGC-13368/2020\|EPI_ISL_712754\|202 | ···················· | ···················· | ···················· | ···················· | ···················· | ···················· |
| X | 6499 hCoV-19/Denmark/DCGC-13369/2020\|EPI_ISL_712755\|202 | ···················· | ···················· | ···················· | ···················· | ···················· | ···················· |
| X | 6500 hCoV-19/Denmark/DCGC-13370/2020\|EPI_ISL_712756\|202 | ···················· | ···················· | ···················· | ···················· | ···················· | ···················· |

| | | | | | | | |
|---|---|---|---|---|---|---|---|
| X | 6501 hCoV-19/Denmark/DCGC-13371/2020\|EPI_ISL_712757\|202 | ···················· | ···················· | ···················· | ···················· | ···················· | ···················· |
| X | 6502 hCoV-19/Denmark/DCGC-13372/2020\|EPI_ISL_712758\|202 | ···················· | ···················· | ···················· | ···················· | ···················· | ···················· |
| X | 6503 hCoV-19/Denmark/DCGC-13373/2020\|EPI_ISL_712759\|202 | ···················· | ···················· | ···················· | ···················· | ···················· | ···················· |
| X | 6504 hCoV-19/Denmark/DCGC-13375/2020\|EPI_ISL_712760\|202 | ···················· | ···················· | ···················· | ···················· | ···················· | ···················· |
| X | 6505 hCoV-19/Denmark/DCGC-13377/2020\|EPI_ISL_712761\|202 | ···················· | ·A·················· | ···················· | ···················· | ···················· | ···················· |
| X | 6506 hCoV-19/Denmark/DCGC-13379/2020\|EPI_ISL_712762\|202 | ···················· | ···················· | ···················· | ···················· | ···················· | ···················· |
| X | 6507 hCoV-19/Denmark/DCGC-13380/2020\|EPI_ISL_712763\|202 | ···················· | ···················· | ···················· | ···················· | ···················· | ···················· |
| X | 6508 hCoV-19/Denmark/DCGC-13381/2020\|EPI_ISL_712764\|202 | ···················· | ·A·················· | ···················· | ···················· | ···················· | ···················· |
| X | 6509 hCoV-19/Denmark/DCGC-13382/2020\|EPI_ISL_712765\|202 | ···················· | ···················· | ···················· | ···················· | ···················· | ···················· |
| X | 6510 hCoV-19/Denmark/DCGC-13384/2020\|EPI_ISL_712766\|202 | ···················· | ···················· | ···················· | ···················· | ···················· | ···················· |
| X | 6511 hCoV-19/Denmark/DCGC-13385/2020\|EPI_ISL_712767\|202 | ···················· | ···················· | ···················· | ···················· | ···················· | ···················· |
| X | 6512 hCoV-19/Denmark/DCGC-13390/2020\|EPI_ISL_712768\|202 | ···················· | ···················· | ···················· | ···················· | ···················· | ···················· |
| X | 6513 hCoV-19/Denmark/DCGC-13391/2020\|EPI_ISL_712769\|202 | ···················· | ···················· | ···················· | ···················· | ···················· | ···················· |
| X | 6514 hCoV-19/Denmark/DCGC-13392/2020\|EPI_ISL_712770\|202 | ···················· | ···················· | ···················· | ···················· | ···················· | ···················· |
| X | 6515 hCoV-19/Denmark/DCGC-13393/2020\|EPI_ISL_712771\|202 | ···················· | ···················· | ···················· | ···················· | ···················· | ···················· |
| X | 6516 hCoV-19/Denmark/DCGC-13396/2020\|EPI_ISL_712772\|202 | ···················· | ···················· | ···················· | ···················· | ···················· | ···················· |
| X | 6517 hCoV-19/Denmark/DCGC-13398/2020\|EPI_ISL_712773\|202 | ···················· | ···················· | ···················· | ···················· | ···················· | ···················· |
| X | 6518 hCoV-19/Denmark/DCGC-13399/2020\|EPI_ISL_712774\|202 | ···················· | ···················· | ···················· | ···················· | ···················· | ···················· |
| X | 6519 hCoV-19/Denmark/DCGC-13400/2020\|EPI_ISL_712775\|202 | ···················· | ···················· | ···················· | ···················· | ···················· | ···················· |
| X | 6520 hCoV-19/Denmark/DCGC-13404/2020\|EPI_ISL_712776\|202 | ···················· | ·A·················· | ···················· | ···················· | ···················· | ···················· |
| X | 6521 hCoV-19/Denmark/DCGC-13405/2020\|EPI_ISL_712777\|202 | ···················· | ···················· | ···················· | ···················· | ···················· | ···················· |
| X | 6522 hCoV-19/Denmark/DCGC-13406/2020\|EPI_ISL_712778\|202 | ···················· | ···················· | ···················· | ···················· | ···················· | ···················· |
| X | 6523 hCoV-19/Denmark/DCGC-13407/2020\|EPI_ISL_712779\|202 | ···················· | ···················· | ···················· | ···················· | ···················· | ···················· |
| X | 6524 hCoV-19/Denmark/DCGC-13423/2020\|EPI_ISL_712790\|202 | ···················· | ···················· | ···················· | ···················· | ······T············· | ···················· |
| X | 6525 hCoV-19/Denmark/DCGC-13425/2020\|EPI_ISL_712791\|202 | ···················· | ···················· | ···················· | ···················· | ···················· | ···················· |
| X | 6526 hCoV-19/Denmark/DCGC-13428/2020\|EPI_ISL_712792\|202 | ···················· | ···················· | ···················· | ···················· | ···················· | ···················· |
| X | 6527 hCoV-19/Denmark/DCGC-13430/2020\|EPI_ISL_712793\|202 | ···················· | ···················· | ···················· | ···················· | ···················· | ···················· |
| X | 6528 hCoV-19/Denmark/DCGC-13431/2020\|EPI_ISL_712794\|202 | ···················· | ···················· | ···················· | ···················· | ···················· | ···················· |
| X | 6529 hCoV-19/Denmark/DCGC-13432/2020\|EPI_ISL_712795\|202 | ···················· | ···················· | ···················· | ···················· | ···················· | ···················· |
| X | 6530 hCoV-19/Denmark/DCGC-13433/2020\|EPI_ISL_712796\|202 | ···················· | ···················· | ···················· | ···················· | ···················· | ···················· |
| X | 6531 hCoV-19/Denmark/DCGC-13434/2020\|EPI_ISL_712797\|202 | ···················· | ···················· | ···················· | ···················· | ···················· | ···················· |
| X | 6532 hCoV-19/Denmark/DCGC-13435/2020\|EPI_ISL_712798\|202 | ···················· | ···················· | ···················· | ···················· | ···················· | ···················· |
| X | 6533 hCoV-19/Denmark/DCGC-13436/2020\|EPI_ISL_712799\|202 | ···················· | ···················· | ···················· | ···················· | ···················· | ···················· |
| X | 6534 hCoV-19/England/ALDP-C49B77/2020\|EPI_ISL_718801\|20 | ···················· | ···················· | ···················· | ···················· | ···················· | ···················· |
| X | 6535 hCoV-19/England/CAMC-C477B1/2020\|EPI_ISL_718805\|20 | ···················· | ···················· | ···················· | ···················· | ···················· | ···················· |
| X | 6536 hCoV-19/England/ALDP-C47E43/2020\|EPI_ISL_718807\|20 | ···················· | ···················· | ···················· | ···················· | ···················· | ···················· |
| X | 6537 hCoV-19/England/ALDP-C4801F/2020\|EPI_ISL_718808\|20 | ···················· | ···················· | ···················· | ···················· | ···················· | ···················· |
| X | 6538 hCoV-19/England/CAMC-C476D2/2020\|EPI_ISL_718814\|20 | ···················· | ···················· | ···················· | ···················· | ···················· | ···················· |
| X | 6539 hCoV-19/England/CAMC-C46EF9/2020\|EPI_ISL_718815\|20 | ···················· | ···················· | ···················· | ···················· | ···················· | ···················· |
| X | 6540 hCoV-19/England/CAMC-C4720B/2020\|EPI_ISL_718816\|20 | ···················· | ···················· | ···················· | ···················· | ······T············· | ···················· |
| X | 6541 hCoV-19/England/CAMC-C4705C/2020\|EPI_ISL_718817\|20 | ···················· | ···················· | ···················· | ···················· | ···················· | ···················· |
| X | 6542 hCoV-19/England/ALDP-C49A98/2020\|EPI_ISL_718819\|20 | ···················· | ···················· | ···················· | ···················· | ···················· | ···················· |
| X | 6543 hCoV-19/England/CAMC-C46D92/2020\|EPI_ISL_718820\|20 | ···················· | ···················· | ···················· | ···················· | ···················· | ···················· |
| X | 6544 hCoV-19/England/ALDP-C49AB6/2020\|EPI_ISL_718827\|20 | ···················· | ···················· | ···················· | ···················· | ···················· | ···················· |
| X | 6545 hCoV-19/England/ALDP-C47EF8/2020\|EPI_ISL_718828\|20 | ···················· | ···················· | ···················· | ···················· | ···················· | ···················· |
| X | 6546 hCoV-19/England/ALDP-C4998C/2020\|EPI_ISL_718829\|20 | ···················· | ···················· | ···················· | ···················· | ···················· | ···················· |
| X | 6547 hCoV-19/England/ALDP-C497A0/2020\|EPI_ISL_718830\|20 | ···················· | ···················· | ···················· | ···················· | ······T············· | ···················· |
| X | 6548 hCoV-19/England/ALDP-C49B59/2020\|EPI_ISL_718831\|20 | ···················· | ···················· | ···················· | ···················· | ···················· | ···················· |
| X | 6549 hCoV-19/England/ALDP-C49AC5/2020\|EPI_ISL_718832\|20 | ···················· | ···················· | ···················· | ···················· | ···················· | ···················· |
| X | 6550 hCoV-19/England/ALDP-C49913/2020\|EPI_ISL_718833\|20 | ···················· | ···················· | ···················· | ···················· | ···················· | ···················· |
| X | 6551 hCoV-19/England/ALDP-C47D55/2020\|EPI_ISL_718837\|20 | ···················· | ····A··············· | ···················· | ···················· | ···················· | ···················· |
| X | 6552 hCoV-19/England/ALDP-C49BA4/2020\|EPI_ISL_718841\|20 | ···················· | ···················· | ···················· | ···················· | ···················· | ···················· |
| X | 6553 hCoV-19/England/CAMC-C4706B/2020\|EPI_ISL_718843\|20 | ···················· | ···················· | ···················· | ···················· | ···················· | ···················· |
| X | 6554 hCoV-19/England/ALDP-C47CD0/2020\|EPI_ISL_718847\|20 | ···················· | ···················· | ···················· | ···················· | ···················· | ···················· |
| X | 6555 hCoV-19/England/ALDP-C48185/2020\|EPI_ISL_718849\|20 | ···················· | ···················· | ···················· | ···················· | ···················· | ···················· |
| X | 6556 hCoV-19/England/ALDP-C481C1/2020\|EPI_ISL_718850\|20 | ···················· | ···················· | ···················· | ···················· | ···················· | ···················· |
| X | 6557 hCoV-19/England/ALDP-C47E70/2020\|EPI_ISL_718853\|20 | ···················· | ···················· | ···················· | ···················· | ···················· | ···················· |
| X | 6558 hCoV-19/England/CAMC-C47308/2020\|EPI_ISL_718857\|20 | ···················· | ···················· | ···················· | ···················· | ···················· | ···················· |
| X | 6559 hCoV-19/England/ALDP-C49AD4/2020\|EPI_ISL_718858\|20 | ···················· | ···················· | ···················· | ···················· | ···················· | ···················· |
| X | 6560 hCoV-19/England/CAMC-C47678/2020\|EPI_ISL_718860\|20 | ···················· | ···················· | ···················· | ···················· | ···················· | ···················· |
| X | 6561 hCoV-19/England/CAMC-C471FF/2020\|EPI_ISL_718863\|20 | ···················· | ···················· | ···················· | ···················· | ···················· | ···················· |
| X | 6562 hCoV-19/England/ALDP-C497DD/2020\|EPI_ISL_718864\|20 | ···················· | ···················· | ···················· | ···················· | ···················· | ···················· |
| X | 6563 hCoV-19/England/ALDP-C49CFC/2020\|EPI_ISL_718866\|20 | ···················· | ···················· | ···················· | ···················· | ···················· | ···················· |
| X | 6564 hCoV-19/England/CAMC-C47432/2020\|EPI_ISL_718867\|20 | ···················· | ···················· | ···················· | ···················· | ······T············· | ···················· |
| X | 6565 hCoV-19/England/CAMC-C47274/2020\|EPI_ISL_718868\|20 | ···················· | ···················· | ···················· | ···················· | ···················· | ···················· |
| X | 6566 hCoV-19/England/ALDP-C4996E/2020\|EPI_ISL_718871\|20 | ···················· | ···················· | ···················· | ···················· | ···················· | ···················· |
| X | 6567 hCoV-19/England/ALDP-C4999B/2020\|EPI_ISL_718872\|20 | ···················· | ···················· | ···················· | ···················· | ···················· | ···················· |
| X | 6568 hCoV-19/England/ALDP-C49825/2020\|EPI_ISL_718873\|20 | ···················· | ···················· | ···················· | ···················· | ···················· | ···················· |
| X | 6569 hCoV-19/England/CAMC-C476A5/2020\|EPI_ISL_718874\|20 | ···················· | ···················· | ···················· | ···················· | ···················· | ···················· |
| X | 6570 hCoV-19/England/CAMC-C46E08/2020\|EPI_ISL_718875\|20 | ···················· | ···················· | ···················· | ···················· | ···················· | ···················· |
| X | 6571 hCoV-19/England/ALDP-C49C83/2020\|EPI_ISL_718877\|20 | ···················· | ···················· | ···················· | ···················· | ···················· | ···················· |
| X | 6572 hCoV-19/England/CAMC-C46E62/2020\|EPI_ISL_718879\|20 | ···················· | ···················· | ···················· | ···················· | ···················· | ···················· |
| X | 6573 hCoV-19/England/CAMC-C46E44/2020\|EPI_ISL_718883\|20 | ···················· | ···················· | ···················· | ···················· | ···················· | ···················· |
| X | 6574 hCoV-19/England/ALDP-C47E07/2020\|EPI_ISL_718887\|20 | ···················· | ···················· | ···················· | ···················· | ···················· | ···················· |
| X | 6575 hCoV-19/England/ALDP-C480F1/2020\|EPI_ISL_718888\|20 | ···················· | ···················· | ···················· | ···················· | ···················· | ···················· |
| X | 6576 hCoV-19/England/ALDP-C49AE3/2020\|EPI_ISL_718889\|20 | ···················· | ···················· | ···················· | ···················· | ···················· | ···················· |
| X | 6577 hCoV-19/England/ALDP-C49AF2/2020\|EPI_ISL_718891\|20 | ···················· | ···················· | ···················· | ···················· | ···················· | ···················· |
| X | 6578 hCoV-19/Brazil/SP-L34-CD881/2020\|EPI_ISL_722129\|20 | ···················· | ······A············· | ···················· | ···················· | ···················· | ···················· |
| X | 6579 hCoV-19/Netherlands/FR-RIVM-20196/2020\|EPI_ISL_723 | ···················· | ···················· | ···················· | ···················· | ···················· | ···················· |
| X | 6580 hCoV-19/Netherlands/FR-RIVM-20197/2020\|EPI_ISL_723 | ···················· | ···················· | ···················· | ···················· | ······T············· | ···················· |

| | | | | | | | | |
|---|---|---|---|---|---|---|---|---|
| X | 6581 | hCoV-19/Netherlands/FR-RIVM-20198/2020\|EPI_ISL_723 | ···················· | ···················· | ···················· | ···················· | ···················· | ···················· |
| X | 6582 | hCoV-19/Netherlands/FR-RIVM-20199/2020\|EPI_ISL_723 | ···················· | ···················· | ···················· | ···················· | ···················· | ···················· |
| X | 6583 | hCoV-19/Netherlands/FR-RIVM-20200/2020\|EPI_ISL_723 | ···················· | ···················· | ···················· | ···················· | ···················· | ···················· |
| X | 6584 | hCoV-19/Netherlands/FR-RIVM-20201/2020\|EPI_ISL_723 | ···················· | ···················· | ···················· | ···················· | ······T············· | ···················· |
| X | 6585 | hCoV-19/Netherlands/FR-RIVM-20203/2020\|EPI_ISL_723 | ···················· | ···················· | ···················· | ···················· | ···················· | ···················· |
| X | 6586 | hCoV-19/Netherlands/FR-RIVM-20204/2020\|EPI_ISL_723 | ···················· | ···················· | ···················· | ···················· | ···················· | ···················· |
| X | 6587 | hCoV-19/Netherlands/FR-RIVM-20205/2020\|EPI_ISL_723 | ···················· | ···················· | ···················· | ···················· | ······T············· | ···················· |
| X | 6588 | hCoV-19/Netherlands/NB-RIVM-20206/2020\|EPI_ISL_723 | ···················· | ···················· | ···················· | ···················· | ···················· | ···················· |
| X | 6589 | hCoV-19/Netherlands/NH-RIVM-20230/2020\|EPI_ISL_723 | ···················· | ···················· | ···················· | ···················· | ···················· | ···················· |
| X | 6590 | hCoV-19/Netherlands/NH-RIVM-20231/2020\|EPI_ISL_723 | ···················· | ···················· | ···················· | ···················· | ···················· | ···················· |
| X | 6591 | hCoV-19/Netherlands/NH-RIVM-20232/2020\|EPI_ISL_723 | ···················· | ···················· | ···················· | ···················· | ···················· | ···················· |
| X | 6592 | hCoV-19/Netherlands/NB-RIVM-20234/2020\|EPI_ISL_723 | ···················· | ···················· | ···················· | ···················· | ···················· | ···················· |
| X | 6593 | hCoV-19/Netherlands/OV-RIVM-20235/2020\|EPI_ISL_723 | ···················· | ···················· | ···················· | ···················· | ···················· | ···················· |
| X | 6594 | hCoV-19/Netherlands/OV-RIVM-20236/2020\|EPI_ISL_723 | ···················· | ···················· | ···················· | ···················· | ···················· | ···················· |
| X | 6595 | hCoV-19/Netherlands/LI-RIVM-20237/2020\|EPI_ISL_723 | ···················· | ···················· | ···················· | ···················· | ···················· | ···················· |
| X | 6596 | hCoV-19/Netherlands/LI-RIVM-20238/2020\|EPI_ISL_723 | ···················· | ·A·················· | ···················· | ···················· | ···················· | ···················· |
| X | 6597 | hCoV-19/Netherlands/LI-RIVM-20239/2020\|EPI_ISL_723 | ···················· | ···················· | ···················· | ···················· | ···················· | ···················· |
| X | 6598 | hCoV-19/Netherlands/LI-RIVM-20240/2020\|EPI_ISL_723 | ···················· | ···················· | ···················· | ···················· | ···················· | ···················· |
| X | 6599 | hCoV-19/Netherlands/LI-RIVM-20242/2020\|EPI_ISL_723 | ···················· | ···················· | ···················· | ···················· | ···················· | ···················· |
| X | 6600 | hCoV-19/Netherlands/LI-RIVM-20243/2020\|EPI_ISL_723 | ···················· | ···················· | ···················· | ···················· | ···················· | ···················· |
| X | 6601 | hCoV-19/Netherlands/LI-RIVM-20245/2020\|EPI_ISL_723 | ···················· | ···················· | ···················· | ···················· | ···················· | ···················· |
| X | 6602 | hCoV-19/Netherlands/LI-RIVM-20246/2020\|EPI_ISL_723 | ···················· | ·A·················· | ···················· | ···················· | ···················· | ···················· |
| X | 6603 | hCoV-19/Netherlands/DR-RIVM-20247/2020\|EPI_ISL_723 | ···················· | ···················· | ···················· | ···················· | ···················· | ···················· |
| X | 6604 | hCoV-19/Netherlands/GR-RIVM-20248/2020\|EPI_ISL_723 | ···················· | ···················· | ···················· | ···················· | ···················· | ···················· |
| X | 6605 | hCoV-19/Netherlands/DR-RIVM-20249/2020\|EPI_ISL_723 | ···················· | ···················· | ···················· | ···················· | ···················· | ···················· |
| X | 6606 | hCoV-19/Netherlands/DR-RIVM-20250/2020\|EPI_ISL_723 | ···················· | ···················· | ···················· | ···················· | ···················· | ···················· |
| X | 6607 | hCoV-19/Netherlands/GR-RIVM-20251/2020\|EPI_ISL_723 | ···················· | ···················· | ···················· | ···················· | ···················· | ···················· |
| X | 6608 | hCoV-19/Netherlands/GR-RIVM-20252/2020\|EPI_ISL_723 | ···················· | ···················· | ···················· | ···················· | ···················· | ···················· |
| X | 6609 | hCoV-19/Netherlands/GR-RIVM-20253/2020\|EPI_ISL_723 | ···················· | ···················· | ···················· | ···················· | ···················· | ···················· |
| X | 6610 | hCoV-19/Netherlands/GR-RIVM-20254/2020\|EPI_ISL_723 | ···················· | ···················· | ···················· | ···················· | ···················· | ···················· |
| X | 6611 | hCoV-19/Netherlands/FR-RIVM-20255/2020\|EPI_ISL_723 | ···················· | ···················· | ···················· | ···················· | ···················· | ···················· |
| X | 6612 | hCoV-19/Netherlands/GR-RIVM-20256/2020\|EPI_ISL_723 | ···················· | ···················· | ···················· | ···················· | ···················· | ···················· |
| X | 6613 | hCoV-19/Netherlands/DR-RIVM-20257/2020\|EPI_ISL_723 | ···················· | ···················· | ···················· | ···················· | ······T············· | ···················· |
| X | 6614 | hCoV-19/Netherlands/GR-RIVM-20258/2020\|EPI_ISL_723 | ···················· | ···················· | ···················· | ···················· | ···················· | ···················· |
| X | 6615 | hCoV-19/Netherlands/NB-RIVM-20259/2020\|EPI_ISL_723 | ···················· | ···················· | ···················· | ···················· | ···················· | ···················· |
| X | 6616 | hCoV-19/Netherlands/GE-RIVM-20260/2020\|EPI_ISL_723 | ···················· | ···················· | ···················· | ···················· | ···················· | ···················· |
| X | 6617 | hCoV-19/Netherlands/NB-RIVM-20261/2020\|EPI_ISL_723 | ···················· | ···················· | ···················· | ···················· | ···················· | ···················· |
| X | 6618 | hCoV-19/Netherlands/NB-RIVM-20263/2020\|EPI_ISL_723 | ···················· | ···················· | ···················· | ···················· | ···················· | ···················· |
| X | 6619 | hCoV-19/Netherlands/NB-RIVM-20266/2020\|EPI_ISL_723 | ···················· | ···················· | ···················· | ···················· | ···················· | ···················· |
| X | 6620 | hCoV-19/Netherlands/NB-RIVM-20268/2020\|EPI_ISL_723 | ···················· | ···················· | ···················· | ···················· | ···················· | ···················· |
| X | 6621 | hCoV-19/Netherlands/NB-RIVM-20269/2020\|EPI_ISL_723 | ···················· | ···················· | ···················· | ···················· | ···················· | ···················· |
| X | 6622 | hCoV-19/Netherlands/NB-RIVM-20270/2020\|EPI_ISL_723 | ···················· | ···················· | ···················· | ···················· | ···················· | ···················· |
| X | 6623 | hCoV-19/Netherlands/NB-RIVM-20272/2020\|EPI_ISL_723 | ···················· | ···················· | ···················· | ···················· | ···················· | ···················· |
| X | 6624 | hCoV-19/Netherlands/NB-RIVM-20273/2020\|EPI_ISL_723 | ···················· | ···················· | ···················· | ···················· | ···················· | ···················· |
| X | 6625 | hCoV-19/Netherlands/NB-RIVM-20276/2020\|EPI_ISL_723 | ···················· | ···················· | ···················· | ···················· | ···················· | ···················· |
| X | 6626 | hCoV-19/Netherlands/NB-RIVM-20277/2020\|EPI_ISL_723 | ···················· | ···················· | ···················· | ···················· | ···················· | ···················· |
| X | 6627 | hCoV-19/Netherlands/NB-RIVM-20278/2020\|EPI_ISL_723 | ···················· | ···················· | ···················· | ···················· | ···················· | ···················· |
| X | 6628 | hCoV-19/Netherlands/GE-RIVM-20280/2020\|EPI_ISL_723 | ···················· | ···················· | ···················· | ···················· | ···················· | ···················· |
| X | 6629 | hCoV-19/Netherlands/NB-RIVM-20281/2020\|EPI_ISL_723 | ···················· | ···················· | ···················· | ···················· | ···················· | ···················· |
| X | 6630 | hCoV-19/Netherlands/NB-RIVM-20282/2020\|EPI_ISL_723 | ···················· | ···················· | ···················· | ···················· | ···················· | ···················· |
| X | 6631 | hCoV-19/Netherlands/GE-RIVM-20283/2020\|EPI_ISL_723 | ···················· | ···················· | ···················· | ···················· | ···················· | ···················· |
| X | 6632 | hCoV-19/Netherlands/GE-RIVM-20285/2020\|EPI_ISL_723 | ···················· | ···················· | ···················· | ···················· | ···················· | ···················· |
| X | 6633 | hCoV-19/Netherlands/GE-RIVM-20286/2020\|EPI_ISL_723 | ···················· | ···················· | ···················· | ···················· | ···················· | ···················· |
| X | 6634 | hCoV-19/Netherlands/GE-RIVM-20287/2020\|EPI_ISL_723 | ···················· | ···················· | ···················· | ···················· | ···················· | ···················· |
| X | 6635 | hCoV-19/Netherlands/GE-RIVM-20288/2020\|EPI_ISL_723 | ···················· | ···················· | ···················· | ···················· | ···················· | ···················· |
| X | 6636 | hCoV-19/Netherlands/GE-RIVM-20300/2020\|EPI_ISL_723 | ···················· | ···················· | ···················· | ···················· | ···················· | ···················· |
| X | 6637 | hCoV-19/Netherlands/FR-RIVM-20301/2020\|EPI_ISL_723 | ···················· | ···················· | ···················· | ···················· | ···················· | ···················· |
| X | 6638 | hCoV-19/Netherlands/FR-RIVM-20302/2020\|EPI_ISL_723 | ···················· | ···················· | ···················· | ···················· | ···················· | ···················· |
| X | 6639 | hCoV-19/Netherlands/FR-RIVM-20303/2020\|EPI_ISL_723 | ···················· | ···················· | ···················· | ···················· | ···················· | ···················· |
| X | 6640 | hCoV-19/Netherlands/FR-RIVM-20304/2020\|EPI_ISL_723 | ···················· | ···················· | ···················· | ···················· | ···················· | ···················· |
| X | 6641 | hCoV-19/Netherlands/FR-RIVM-20305/2020\|EPI_ISL_723 | ···················· | ···················· | ···················· | ···················· | ···················· | ···················· |
| X | 6642 | hCoV-19/Netherlands/FR-RIVM-20306/2020\|EPI_ISL_723 | ···················· | ···················· | ···················· | ···················· | ···················· | ···················· |
| X | 6643 | hCoV-19/Netherlands/GE-RIVM-20307/2020\|EPI_ISL_723 | ···················· | ···················· | ···················· | ···················· | ···················· | ···················· |
| X | 6644 | hCoV-19/Netherlands/LI-RIVM-20308/2020\|EPI_ISL_723 | ···················· | ···················· | ···················· | ···················· | ···················· | ···················· |
| X | 6645 | hCoV-19/Netherlands/GE-RIVM-20309/2020\|EPI_ISL_723 | ···················· | ···················· | ···················· | ···················· | ···················· | ···················· |
| X | 6646 | hCoV-19/Netherlands/GE-RIVM-20310/2020\|EPI_ISL_723 | ···················· | ···················· | ···················· | ···················· | ···················· | ···················· |
| X | 6647 | hCoV-19/Netherlands/GE-RIVM-20311/2020\|EPI_ISL_723 | ···················· | ···················· | ···················· | ···················· | ···················· | ···················· |
| X | 6648 | hCoV-19/Netherlands/GE-RIVM-20312/2020\|EPI_ISL_723 | ···················· | ···················· | ···················· | ···················· | ···················· | ···················· |
| X | 6649 | hCoV-19/Netherlands/OV-RIVM-20313/2020\|EPI_ISL_723 | ···················· | ···················· | ···················· | ···················· | ···················· | ···················· |
| X | 6650 | hCoV-19/Netherlands/OV-RIVM-20315/2020\|EPI_ISL_723 | ···················· | ···················· | ···················· | ···················· | ···················· | ···················· |
| X | 6651 | hCoV-19/Netherlands/FR-RIVM-20316/2020\|EPI_ISL_723 | ···················· | ···················· | ···················· | ···················· | ···················· | ···················· |
| X | 6652 | hCoV-19/Netherlands/GE-RIVM-20321/2020\|EPI_ISL_723 | ···················· | ···················· | ···················· | ···················· | ···················· | ···················· |
| X | 6653 | hCoV-19/Netherlands/GE-RIVM-20322/2020\|EPI_ISL_723 | ···················· | ···················· | ···················· | ···················· | ···················· | ···················· |
| X | 6654 | hCoV-19/Netherlands/GE-RIVM-20323/2020\|EPI_ISL_723 | ···················· | ···················· | ···················· | ···················· | ···················· | ···················· |
| X | 6655 | hCoV-19/Netherlands/GE-RIVM-20327/2020\|EPI_ISL_723 | ···················· | ···················· | ···················· | ···················· | ···················· | ···················· |
| X | 6656 | hCoV-19/Netherlands/GE-RIVM-20329/2020\|EPI_ISL_723 | ···················· | ···················· | ···················· | ···················· | ···················· | ···················· |
| X | 6657 | hCoV-19/Denmark/DCGC-13916/2020\|EPI_ISL_711449\|202 | ···················· | ···················· | ···················· | ···················· | ···················· | ···················· |
| X | 6658 | hCoV-19/Denmark/DCGC-13917/2020\|EPI_ISL_711450\|202 | ···················· | ···················· | ···················· | ···················· | ···················· | ···················· |
| X | 6659 | hCoV-19/Denmark/DCGC-13918/2020\|EPI_ISL_711451\|202 | ···················· | ···················· | ···················· | ···················· | ···················· | ···················· |
| X | 6660 | hCoV-19/Denmark/DCGC-13920/2020\|EPI_ISL_711452\|202 | ···················· | ···················· | ···················· | ···················· | ···················· | ···················· |

| | | | | | | | |
|---|---|---|---|---|---|---|---|
| X | 6661 hCoV-19/Denmark/DCGC-13921/2020\|EPI_ISL_711453\|202 | ···················· | ···················· | ···················· | ···················· | ···················· | ···················· |
| X | 6662 hCoV-19/Denmark/DCGC-13922/2020\|EPI_ISL_711454\|202 | ···················· | ···················· | ···················· | ···················· | ···················· | ···················· |
| X | 6663 hCoV-19/Denmark/DCGC-13924/2020\|EPI_ISL_711455\|202 | ···················· | ···················· | ···················· | ···················· | ···················· | ···················· |
| X | 6664 hCoV-19/Denmark/DCGC-13925/2020\|EPI_ISL_711456\|202 | ···················· | ···················· | ···················· | ···················· | ···················· | ···················· |
| X | 6665 hCoV-19/Denmark/DCGC-13926/2020\|EPI_ISL_711457\|202 | ···················· | ···················· | ···················· | ···················· | ···················· | ···················· |
| X | 6666 hCoV-19/Denmark/DCGC-13927/2020\|EPI_ISL_711458\|202 | ···················· | ···················· | ···················· | ···················· | ···················· | ···················· |
| X | 6667 hCoV-19/Denmark/DCGC-13928/2020\|EPI_ISL_711459\|202 | ···················· | ···················· | ···················· | ···················· | ···················· | ···················· |
| X | 6668 hCoV-19/Denmark/DCGC-13964/2020\|EPI_ISL_711480\|202 | ···················· | ·A·················· | ···················· | ···················· | ···················· | ···················· |
| X | 6669 hCoV-19/Denmark/DCGC-13965/2020\|EPI_ISL_711481\|202 | ···················· | ·A·················· | ···················· | ···················· | ···················· | ··A················· |
| X | 6670 hCoV-19/Denmark/DCGC-13967/2020\|EPI_ISL_711482\|202 | ···················· | ···················· | ···················· | ···················· | ···················· | ···················· |
| X | 6671 hCoV-19/Denmark/DCGC-13969/2020\|EPI_ISL_711483\|202 | ···················· | ···················· | ···················· | ···················· | ···················· | ···················· |
| X | 6672 hCoV-19/Denmark/DCGC-13970/2020\|EPI_ISL_711484\|202 | ···················· | ···················· | ···················· | ···················· | ···················· | ···················· |
| X | 6673 hCoV-19/Denmark/DCGC-13971/2020\|EPI_ISL_711485\|202 | ···················· | ···················· | ···················· | ···················· | ···················· | ···················· |
| X | 6674 hCoV-19/Denmark/DCGC-13972/2020\|EPI_ISL_711486\|202 | ···················· | ···················· | ···················· | ···················· | ···················· | ···················· |
| X | 6675 hCoV-19/Denmark/DCGC-13974/2020\|EPI_ISL_711487\|202 | ···················· | ···················· | ···················· | ···················· | ···················· | ···················· |
| X | 6676 hCoV-19/Denmark/DCGC-13975/2020\|EPI_ISL_711488\|202 | ···················· | ···················· | ···················· | ···················· | ···················· | ···················· |
| X | 6677 hCoV-19/Denmark/DCGC-13976/2020\|EPI_ISL_711489\|202 | ···················· | ···················· | ···················· | ···················· | ···················· | ···················· |
| X | 6678 hCoV-19/Denmark/DCGC-13977/2020\|EPI_ISL_711490\|202 | ···················· | ···················· | ···················· | ···················· | ···················· | ···················· |
| X | 6679 hCoV-19/Denmark/DCGC-13978/2020\|EPI_ISL_711491\|202 | ··········C········· | ···················· | ···················· | ···················· | ···················· | ···················· |
| X | 6680 hCoV-19/Denmark/DCGC-13980/2020\|EPI_ISL_711492\|202 | ···················· | ···················· | ···················· | ···················· | ···················· | ···················· |
| X | 6681 hCoV-19/Denmark/DCGC-13981/2020\|EPI_ISL_711493\|202 | ··········C········· | ···················· | ···················· | ···················· | ···················· | ···················· |
| X | 6682 hCoV-19/Denmark/DCGC-13983/2020\|EPI_ISL_711494\|202 | ···················· | ···················· | ···················· | ···················· | ···················· | ···················· |
| X | 6683 hCoV-19/Denmark/DCGC-13984/2020\|EPI_ISL_711495\|202 | ···················· | ···················· | ···················· | ···················· | ···················· | ···················· |
| X | 6684 hCoV-19/Denmark/DCGC-13985/2020\|EPI_ISL_711496\|202 | ···················· | ···················· | ···················· | ···················· | ···················· | ···················· |
| X | 6685 hCoV-19/Denmark/DCGC-13986/2020\|EPI_ISL_711497\|202 | ···················· | ···················· | ···················· | ···················· | ···················· | ···················· |
| X | 6686 hCoV-19/Denmark/DCGC-13987/2020\|EPI_ISL_711498\|202 | ···················· | ···················· | ···················· | ···················· | ···················· | ···················· |
| X | 6687 hCoV-19/Denmark/DCGC-13989/2020\|EPI_ISL_711499\|202 | ···················· | ···················· | ···················· | ···················· | ···················· | ···················· |
| X | 6688 hCoV-19/Denmark/DCGC-13991/2020\|EPI_ISL_711500\|202 | ···················· | ···················· | ···················· | ···················· | ···················· | ···················· |
| X | 6689 hCoV-19/Denmark/DCGC-13993/2020\|EPI_ISL_711501\|202 | ···················· | ···················· | ···················· | ···················· | ···················· | ···················· |
| X | 6690 hCoV-19/Denmark/DCGC-13995/2020\|EPI_ISL_711502\|202 | ···················· | ···················· | ···················· | ···················· | ···················· | ···················· |
| X | 6691 hCoV-19/Denmark/DCGC-13996/2020\|EPI_ISL_711503\|202 | ···················· | ···················· | ···················· | ···················· | ···················· | ···················· |
| X | 6692 hCoV-19/Denmark/DCGC-13997/2020\|EPI_ISL_711504\|202 | ···················· | ···················· | ···················· | ···················· | ···················· | ···················· |
| X | 6693 hCoV-19/Denmark/DCGC-13998/2020\|EPI_ISL_711505\|202 | ···················· | ···················· | ···················· | ···················· | ···················· | ···················· |
| X | 6694 hCoV-19/Denmark/DCGC-13999/2020\|EPI_ISL_711506\|202 | ···················· | ···················· | ···················· | ···················· | ···················· | ···················· |
| X | 6695 hCoV-19/Denmark/DCGC-14000/2020\|EPI_ISL_711507\|202 | ···················· | ···················· | ···················· | ···················· | ···················· | ···················· |
| X | 6696 hCoV-19/Denmark/DCGC-14001/2020\|EPI_ISL_711508\|202 | ···················· | ···················· | ···················· | ···················· | ···················· | ···················· |
| X | 6697 hCoV-19/Denmark/DCGC-14002/2020\|EPI_ISL_711509\|202 | ···················· | ···················· | ···················· | ···················· | ···················· | ···················· |
| X | 6698 hCoV-19/Denmark/DCGC-14003/2020\|EPI_ISL_711510\|202 | ···················· | ···················· | ···················· | ···················· | ···················· | ···················· |
| X | 6699 hCoV-19/Denmark/DCGC-14004/2020\|EPI_ISL_711511\|202 | ···················· | ···················· | ···················· | ···················· | ···················· | ···················· |
| X | 6700 hCoV-19/Denmark/DCGC-14005/2020\|EPI_ISL_711512\|202 | ···················· | ···················· | ···················· | ···················· | ···················· | ···················· |
| X | 6701 hCoV-19/Denmark/DCGC-14008/2020\|EPI_ISL_711513\|202 | ···················· | ···················· | ···················· | ···················· | ···················· | ···················· |
| X | 6702 hCoV-19/Denmark/DCGC-14010/2020\|EPI_ISL_711514\|202 | ···················· | ···················· | ···················· | ···················· | ···················· | ···················· |
| X | 6703 hCoV-19/Denmark/DCGC-14053/2020\|EPI_ISL_711537\|202 | ···················· | ···················· | ···················· | ···················· | ···················· | ···················· |
| X | 6704 hCoV-19/Denmark/DCGC-14054/2020\|EPI_ISL_711538\|202 | ···················· | ···················· | ···················· | ···················· | ···················· | ···················· |
| X | 6705 hCoV-19/Denmark/DCGC-14055/2020\|EPI_ISL_711539\|202 | ···················· | ···················· | ···················· | ···················· | ···················· | ···················· |
| X | 6706 hCoV-19/Denmark/DCGC-14066/2020\|EPI_ISL_711546\|202 | ···················· | ···················· | ···················· | ···················· | ···················· | ···················· |
| X | 6707 hCoV-19/Denmark/DCGC-14067/2020\|EPI_ISL_711547\|202 | ···················· | ···················· | ···················· | ···················· | ···················· | ···················· |
| X | 6708 hCoV-19/Denmark/DCGC-13409/2020\|EPI_ISL_712780\|202 | ···················· | ···················· | ···················· | ···················· | ···················· | ···················· |
| X | 6709 hCoV-19/Denmark/DCGC-13411/2020\|EPI_ISL_712781\|202 | ···················· | ···················· | ···················· | ···················· | ···················· | ···················· |
| X | 6710 hCoV-19/Denmark/DCGC-13412/2020\|EPI_ISL_712782\|202 | ···················· | ···················· | ···················· | ···················· | ···················· | ···················· |
| X | 6711 hCoV-19/Denmark/DCGC-13413/2020\|EPI_ISL_712783\|202 | ···················· | ···················· | ···················· | ···················· | ···················· | ···················· |
| X | 6712 hCoV-19/Denmark/DCGC-13414/2020\|EPI_ISL_712784\|202 | ···················· | ···················· | ···················· | ···················· | ···················· | ···················· |
| X | 6713 hCoV-19/Denmark/DCGC-13415/2020\|EPI_ISL_712785\|202 | ···················· | ···················· | ···················· | ···················· | ···················· | ···················· |
| X | 6714 hCoV-19/Denmark/DCGC-13416/2020\|EPI_ISL_712786\|202 | ···················· | ···················· | ···················· | ···················· | ···················· | ···················· |
| X | 6715 hCoV-19/Denmark/DCGC-13417/2020\|EPI_ISL_712787\|202 | ···················· | ···················· | ···················· | ···················· | ···················· | ···················· |
| X | 6716 hCoV-19/Denmark/DCGC-13419/2020\|EPI_ISL_712788\|202 | ···················· | ···················· | ···················· | ···················· | ···················· | ···················· |
| X | 6717 hCoV-19/Denmark/DCGC-13420/2020\|EPI_ISL_712789\|202 | ···················· | ···················· | ···················· | ···················· | ···················· | ···················· |
| X | 6718 hCoV-19/Denmark/DCGC-13437/2020\|EPI_ISL_712800\|202 | ···················· | ···················· | ···················· | ···················· | ···················· | ···················· |
| X | 6719 hCoV-19/Denmark/DCGC-13439/2020\|EPI_ISL_712801\|202 | ···················· | ···················· | ···················· | ···················· | ···················· | ···················· |
| X | 6720 hCoV-19/Denmark/DCGC-13441/2020\|EPI_ISL_712802\|202 | ···················· | ···················· | ···················· | ···················· | ···················· | ···················· |
| X | 6721 hCoV-19/Denmark/DCGC-13442/2020\|EPI_ISL_712803\|202 | ···················· | ···················· | ···················· | ···················· | ···················· | ···················· |
| X | 6722 hCoV-19/Denmark/DCGC-13445/2020\|EPI_ISL_712804\|202 | ···················· | ···················· | ···················· | ···················· | ···················· | ···················· |
| X | 6723 hCoV-19/Denmark/DCGC-13447/2020\|EPI_ISL_712805\|202 | ···················· | ···················· | ···················· | ···················· | ···················· | ···················· |
| X | 6724 hCoV-19/Denmark/DCGC-13450/2020\|EPI_ISL_712806\|202 | ···················· | ···················· | ···················· | ···················· | ···················· | ···················· |
| X | 6725 hCoV-19/Denmark/DCGC-13451/2020\|EPI_ISL_712807\|202 | ···················· | ···················· | ···················· | ···················· | ···················· | ···················· |
| X | 6726 hCoV-19/Denmark/DCGC-13452/2020\|EPI_ISL_712808\|202 | ···················· | ···················· | ···················· | ···················· | ···················· | ···················· |
| X | 6727 hCoV-19/Denmark/DCGC-13454/2020\|EPI_ISL_712809\|202 | ···················· | ···················· | ···················· | ···················· | ···················· | ···················· |
| X | 6728 hCoV-19/Denmark/DCGC-13455/2020\|EPI_ISL_712810\|202 | ···················· | ···················· | ···················· | ···················· | ···················· | ···················· |
| X | 6729 hCoV-19/Denmark/DCGC-13458/2020\|EPI_ISL_712811\|202 | ···················· | ···················· | ···················· | ···················· | ···················· | ···················· |
| X | 6730 hCoV-19/Denmark/DCGC-13459/2020\|EPI_ISL_712812\|202 | ···················· | ···················· | ···················· | ···················· | ···················· | ···················· |
| X | 6731 hCoV-19/Denmark/DCGC-13462/2020\|EPI_ISL_712813\|202 | ···················· | ···················· | ···················· | ···················· | ···················· | ···················· |
| X | 6732 hCoV-19/Denmark/DCGC-13463/2020\|EPI_ISL_712814\|202 | ···················· | ···················· | ···················· | ···················· | ···················· | ···················· |
| X | 6733 hCoV-19/Denmark/DCGC-13464/2020\|EPI_ISL_712815\|202 | ···················· | ···················· | ···················· | ···················· | ···················· | ···················· |
| X | 6734 hCoV-19/Denmark/DCGC-13466/2020\|EPI_ISL_712816\|202 | ···················· | ···················· | ···················· | ···················· | ···················· | ···················· |
| X | 6735 hCoV-19/Denmark/DCGC-13469/2020\|EPI_ISL_712817\|202 | ···················· | ···················· | ···················· | ···················· | ···················· | ···················· |
| X | 6736 hCoV-19/Denmark/DCGC-13471/2020\|EPI_ISL_712818\|202 | ···················· | ···················· | ···················· | ···················· | ···················· | ···················· |
| X | 6737 hCoV-19/Denmark/DCGC-13473/2020\|EPI_ISL_712819\|202 | ···················· | ···················· | ···················· | ···················· | ···················· | ···················· |
| X | 6738 hCoV-19/Denmark/DCGC-13474/2020\|EPI_ISL_712820\|202 | ···················· | ···················· | ···················· | ···················· | ···················· | ···················· |
| X | 6739 hCoV-19/Denmark/DCGC-13476/2020\|EPI_ISL_712821\|202 | ···················· | ···················· | ···················· | ···················· | ···················· | ···················· |
| X | 6740 hCoV-19/Denmark/DCGC-13478/2020\|EPI_ISL_712822\|202 | ···················· | ···················· | ···················· | ···················· | ···················· | ···················· |

| | | | | | | | | |
|---|---|---|---|---|---|---|---|---|
| X | 6741 | hCoV-19/Denmark/DCGC-13480/2020\|EPI_ISL_712823\|202 | ···················· | ···················· | ···················· | ···················· | ···················· | ···················· |
| X | 6742 | hCoV-19/Denmark/DCGC-13485/2020\|EPI_ISL_712824\|202 | ···················· | ···················· | ···················· | ···················· | ···················· | ···················· |
| X | 6743 | hCoV-19/Denmark/DCGC-13486/2020\|EPI_ISL_712825\|202 | ···················· | ···················· | ···················· | ···················· | ···················· | ···················· |
| X | 6744 | hCoV-19/Denmark/DCGC-13487/2020\|EPI_ISL_712826\|202 | ···················· | ···················· | ···················· | ···················· | ···················· | ···················· |
| X | 6745 | hCoV-19/Denmark/DCGC-13490/2020\|EPI_ISL_712827\|202 | ···················· | ···················· | ···················· | ···················· | ···················· | ···················· |
| X | 6746 | hCoV-19/Denmark/DCGC-13491/2020\|EPI_ISL_712828\|202 | ···················· | ···················· | ···················· | ···················· | ···················· | ···················· |
| X | 6747 | hCoV-19/Denmark/DCGC-13492/2020\|EPI_ISL_712829\|202 | ···················· | ···················· | ···················· | ···················· | ···················· | ···················· |
| X | 6748 | hCoV-19/Denmark/DCGC-13493/2020\|EPI_ISL_712830\|202 | ···················· | ···················· | ···················· | ···················· | ···················· | ···················· |
| X | 6749 | hCoV-19/Denmark/DCGC-13495/2020\|EPI_ISL_712831\|202 | ···················· | ···················· | ···················· | ···················· | ···················· | ···················· |
| X | 6750 | hCoV-19/Denmark/DCGC-13496/2020\|EPI_ISL_712832\|202 | ···················· | ···················· | ···················· | ···················· | ···················· | ···················· |
| X | 6751 | hCoV-19/Denmark/DCGC-13497/2020\|EPI_ISL_712833\|202 | ···················· | ···················· | ···················· | ···················· | ···················· | ···················· |
| X | 6752 | hCoV-19/Denmark/DCGC-13498/2020\|EPI_ISL_712834\|202 | ···················· | ···················· | ···················· | ···················· | ···················· | ···················· |
| X | 6753 | hCoV-19/Denmark/DCGC-13499/2020\|EPI_ISL_712835\|202 | ···················· | ···················· | ···················· | ···················· | ···················· | ···················· |
| X | 6754 | hCoV-19/Denmark/DCGC-13500/2020\|EPI_ISL_712836\|202 | ···················· | ···················· | ···················· | ···················· | ···················· | ···················· |
| X | 6755 | hCoV-19/Denmark/DCGC-13501/2020\|EPI_ISL_712837\|202 | ···················· | ···················· | ···················· | ···················· | ···················· | ···················· |
| X | 6756 | hCoV-19/Denmark/DCGC-13502/2020\|EPI_ISL_712838\|202 | ···················· | ···················· | ···················· | ···················· | ···················· | ···················· |
| X | 6757 | hCoV-19/Denmark/DCGC-13503/2020\|EPI_ISL_712839\|202 | ···················· | ···················· | ···················· | ···················· | ···················· | ···················· |
| X | 6758 | hCoV-19/Denmark/DCGC-13504/2020\|EPI_ISL_712840\|202 | ···················· | ···················· | ···················· | ···················· | ···················· | ···················· |
| X | 6759 | hCoV-19/Denmark/DCGC-13505/2020\|EPI_ISL_712841\|202 | ···················· | ···················· | ···················· | ···················· | ···················· | ···················· |
| X | 6760 | hCoV-19/Denmark/DCGC-13506/2020\|EPI_ISL_712842\|202 | ···················· | ···················· | ···················· | ···················· | ···················· | ···················· |
| X | 6761 | hCoV-19/Denmark/DCGC-13507/2020\|EPI_ISL_712843\|202 | ···················· | ···················· | ···················· | ···················· | ···················· | ···················· |
| X | 6762 | hCoV-19/Denmark/DCGC-13509/2020\|EPI_ISL_712844\|202 | ···················· | ···················· | ···················· | ···················· | ···················· | ···················· |
| X | 6763 | hCoV-19/Denmark/DCGC-13510/2020\|EPI_ISL_712845\|202 | ···················· | ···················· | ···················· | ···················· | ···················· | ···················· |
| X | 6764 | hCoV-19/Denmark/DCGC-13542/2020\|EPI_ISL_712868\|202 | ···················· | ···················· | ···················· | ···················· | ···················· | ···················· |
| X | 6765 | hCoV-19/Denmark/DCGC-13544/2020\|EPI_ISL_712869\|202 | ···················· | ···················· | ···················· | ···················· | ···················· | ···················· |
| X | 6766 | hCoV-19/Denmark/DCGC-13555/2020\|EPI_ISL_712876\|202 | ···················· | ·A·················· | ···················· | ···················· | ···················· | ···················· |
| X | 6767 | hCoV-19/Denmark/DCGC-13559/2020\|EPI_ISL_712877\|202 | ···················· | ·A·················· | ···················· | ···················· | ···················· | ···················· |
| X | 6768 | hCoV-19/Denmark/DCGC-13560/2020\|EPI_ISL_712878\|202 | ···················· | ···················· | ···················· | ···················· | ···················· | ···················· |
| X | 6769 | hCoV-19/Belgium/UGent-214/2020\|EPI_ISL_717603\|2020 | ···················· | ···················· | ···················· | ···················· | ···················· | ···················· |
| X | 6778 | hCoV-19/Belgium/UGent-211/2020\|EPI_ISL_717612\|2020 | ···················· | ···················· | ···················· | ···················· | ···················· | ···················· |
| X | 6779 | hCoV-19/Belgium/UGent-212/2020\|EPI_ISL_717613\|2020 | ···················· | ···················· | ···················· | ···················· | ···················· | ···················· |
| X | 6780 | hCoV-19/Belgium/UGent-213/2020\|EPI_ISL_717614\|2020 | ···················· | ···················· | ···················· | ···················· | ···················· | ···················· |
| X | 6786 | hCoV-19/Belgium/UGent-222/2020\|EPI_ISL_717620\|2020 | ···················· | ···················· | ···················· | ···················· | ···················· | ···················· |
| X | 6787 | hCoV-19/Belgium/UGent-223/2020\|EPI_ISL_717621\|2020 | ···················· | ···················· | ···················· | ···················· | ···················· | ···················· |
| X | 6788 | hCoV-19/Belgium/UGent-227/2020\|EPI_ISL_717622\|2020 | ···················· | ···················· | ···················· | ···················· | ···················· | ···················· |
| X | 6789 | hCoV-19/Belgium/UGent-230/2020\|EPI_ISL_717623\|2020 | ···················· | ···················· | ···················· | ···················· | ···················· | ···················· |
| X | 6790 | hCoV-19/Belgium/UGent-231/2020\|EPI_ISL_717624\|2020 | ···················· | ···················· | ···················· | ···················· | ···················· | ···················· |
| X | 6791 | hCoV-19/Belgium/UGent-235/2020\|EPI_ISL_717625\|2020 | ···················· | ···················· | ···················· | ···················· | ···················· | ···················· |
| X | 6792 | hCoV-19/Belgium/UGent-240/2020\|EPI_ISL_717626\|2020 | ···················· | ···················· | ···················· | ···················· | ···················· | ···················· |
| X | 6793 | hCoV-19/Belgium/UGent-242/2020\|EPI_ISL_717627\|2020 | ···················· | ···················· | ···················· | ···················· | ···················· | ···················· |
| X | 6794 | hCoV-19/Belgium/UGent-243/2020\|EPI_ISL_717628\|2020 | ···················· | ···················· | ···················· | ···················· | ···················· | ···················· |
| X | 6795 | hCoV-19/Belgium/UGent-245/2020\|EPI_ISL_717629\|2020 | ···················· | ···················· | ···················· | ···················· | ···················· | ···················· |
| X | 6796 | hCoV-19/Belgium/UGent-246/2020\|EPI_ISL_717630\|2020 | ···················· | ···················· | ···················· | ···················· | ···················· | ···················· |
| X | 6797 | hCoV-19/Belgium/UGent-247/2020\|EPI_ISL_717631\|2020 | ···················· | ···················· | ···················· | ···················· | ···················· | ···················· |
| X | 6798 | hCoV-19/Belgium/UGent-248/2020\|EPI_ISL_717632\|2020 | ···················· | ···················· | ···················· | ···················· | ···················· | ···················· |
| X | 6799 | hCoV-19/Belgium/UGent-249/2020\|EPI_ISL_717633\|2020 | ···················· | ···················· | ···················· | ···················· | ···················· | ···················· |
| X | 6800 | hCoV-19/England/ALDP-C499AA/2020\|EPI_ISL_718902\|20 | ···················· | ···················· | ···················· | ···················· | ···················· | ···················· |
| X | 6801 | hCoV-19/England/ALDP-C498E9/2020\|EPI_ISL_718908\|20 | ···················· | ···················· | ···················· | ···················· | ······T············· | ···················· |
| X | 6802 | hCoV-19/England/ALDP-C48255/2020\|EPI_ISL_718911\|20 | ···················· | ···················· | ···················· | ···················· | ···················· | ···················· |
| X | 6803 | hCoV-19/England/ALDP-C47D82/2020\|EPI_ISL_718914\|20 | ···················· | ···················· | ···················· | ···················· | ···················· | ···················· |
| X | 6804 | hCoV-19/England/ALDP-C49B4A/2020\|EPI_ISL_718915\|20 | ···················· | ······A············· | ···················· | ···················· | ···················· | ···················· |
| X | 6805 | hCoV-19/England/CAMC-C477EE/2020\|EPI_ISL_718918\|20 | ···················· | ···················· | ···················· | ···················· | ···················· | ···················· |
| X | 6806 | hCoV-19/England/ALDP-C49B68/2020\|EPI_ISL_718922\|20 | ···················· | ···················· | ···················· | ···················· | ···················· | ···················· |
| X | 6807 | hCoV-19/England/ALDP-C49BE0/2020\|EPI_ISL_718923\|20 | ···················· | ···················· | ···················· | ···················· | ···················· | ···················· |
| X | 6808 | hCoV-19/England/CAMC-C4703E/2020\|EPI_ISL_718924\|20 | ···················· | ···················· | ···················· | ···················· | ···················· | ···················· |
| X | 6809 | hCoV-19/England/ALDP-C480E2/2020\|EPI_ISL_718929\|20 | ···················· | ···················· | ···················· | ···················· | ···················· | ···················· |
| X | 6810 | hCoV-19/England/ALDP-C49B95/2020\|EPI_ISL_718932\|20 | ···················· | ···················· | ···················· | ···················· | ···················· | ···················· |
| X | 6811 | hCoV-19/England/ALDP-C47FE6/2020\|EPI_ISL_718935\|20 | ···················· | ···················· | ···················· | ···················· | ···················· | ···················· |
| X | 6812 | hCoV-19/England/ALDP-C47DA0/2020\|EPI_ISL_718936\|20 | ···················· | ···················· | ···················· | ···················· | ···················· | ···················· |
| X | 6813 | hCoV-19/England/CAMC-C47809/2020\|EPI_ISL_718937\|20 | ···················· | ···················· | ···················· | ···················· | ···················· | ···················· |
| X | 6814 | hCoV-19/England/CAMC-C470E3/2020\|EPI_ISL_718938\|20 | ···················· | ···················· | ···················· | ···················· | ···················· | ···················· |
| X | 6815 | hCoV-19/England/CAMC-C47520/2020\|EPI_ISL_718940\|20 | ···················· | ···················· | ···················· | ···················· | ···················· | ···················· |
| X | 6816 | hCoV-19/England/ALDP-C49BFF/2020\|EPI_ISL_718943\|20 | ···················· | ···················· | ···················· | ···················· | ···················· | ···················· |
| X | 6817 | hCoV-19/England/ALDP-C49904/2020\|EPI_ISL_718945\|20 | ···················· | ···················· | ···················· | ···················· | ···················· | ···················· |
| X | 6818 | hCoV-19/England/CAMC-C4710E/2020\|EPI_ISL_718948\|20 | ···················· | ···················· | ···················· | ···················· | ···················· | ···················· |
| X | 6819 | hCoV-19/England/ALDP-C47CEF/2020\|EPI_ISL_718950\|20 | ···················· | ···················· | ···················· | ···················· | ···················· | ···················· |
| X | 6820 | hCoV-19/England/ALDP-C47C94/2020\|EPI_ISL_718952\|20 | ···················· | ···················· | ···················· | ···················· | ···················· | ···················· |
| X | 6821 | hCoV-19/England/ALDP-C497CE/2020\|EPI_ISL_718953\|20 | ···················· | ···················· | ···················· | ···················· | ···················· | ···················· |
| X | 6822 | hCoV-19/England/CAMC-C47906/2020\|EPI_ISL_718954\|20 | ···················· | ····A··············· | ···················· | ···················· | ···················· | ···················· |
| X | 6823 | hCoV-19/England/ALDP-C4813A/2020\|EPI_ISL_718957\|20 | ···················· | ···················· | ···················· | ···················· | ···················· | ···················· |
| X | 6824 | hCoV-19/England/CAMC-C476E1/2020\|EPI_ISL_718958\|20 | ···················· | ···················· | ···················· | ···················· | ···················· | ···················· |
| X | 6825 | hCoV-19/England/ALDP-C4989E/2020\|EPI_ISL_718962\|20 | ···················· | ···················· | ···················· | ···················· | ···················· | ···················· |
| X | 6826 | hCoV-19/England/ALDP-C47F5F/2020\|EPI_ISL_718963\|20 | ···················· | ···················· | ···················· | ···················· | ···················· | ···················· |
| X | 6827 | hCoV-19/England/ALDP-C47CFE/2020\|EPI_ISL_718964\|20 | ···················· | ···················· | ···················· | ···················· | ···················· | ···················· |
| X | 6828 | hCoV-19/England/ALDP-C47F13/2020\|EPI_ISL_718966\|20 | ···················· | ···················· | ···················· | ···················· | ···················· | ···················· |
| X | 6829 | hCoV-19/England/ALDP-C49B1D/2020\|EPI_ISL_718970\|20 | ···················· | ···················· | ···················· | ···················· | ···················· | ···················· |
| X | 6830 | hCoV-19/England/ALDP-C47D46/2020\|EPI_ISL_718977\|20 | ···················· | ····A··············· | ···················· | ···················· | ···················· | ···················· |
| X | 6831 | hCoV-19/England/ALDP-C480C4/2020\|EPI_ISL_718978\|20 | ···················· | ···················· | ···················· | ···················· | ···················· | ···················· |
| X | 6832 | hCoV-19/England/ALDP-C49922/2020\|EPI_ISL_718979\|20 | ···················· | ·A·················· | ···················· | ···················· | ···················· | ···A················ |
| X | 6833 | hCoV-19/England/CAMC-C40AFB/2020\|EPI_ISL_718981\|20 | ···················· | ·A·················· | ···················· | ···················· | ···················· | ···················· |

```
X 6834 hCoV-19/England/CAMC-C42482/2020|EPI_ISL_718985|20 ···················· ···················· ···················· ···················· ···················· ····················
X 6835 hCoV-19/England/CAMC-C3EE12/2020|EPI_ISL_718986|20 ···················· ···················· ···················· ···················· ···················· ····················
X 6836 hCoV-19/England/CAMC-C4226A/2020|EPI_ISL_718997|20 ···················· ···················· ···················· ···················· ···················· ····················
X 6837 hCoV-19/England/CAMC-C44F16/2020|EPI_ISL_718998|20 ···················· ···················· ···················· ···················· ······T············· ····················
X 6838 hCoV-19/England/CAMC-C3F048/2020|EPI_ISL_718999|20 ···················· ···················· ···················· ···················· ···················· ····················
X 6839 hCoV-19/India/MH-NCCS-NR-14432/2020|EPI_ISL_722177 ···················· ···················· ···················· ···················· ···················· ····················
X 6840 hCoV-19/India/MH-NCCS-NR-8740/2020|EPI_ISL_722178| ···················· ···················· ···················· ···················· ···················· ····················
X 6841 hCoV-19/India/MH-NCCS-NR-15431/2020|EPI_ISL_722179 ···················· ···················· ···················· ···················· ···················· ····················
X 6842 hCoV-19/India/MH-NCCS-NR-12775/2020|EPI_ISL_722180 ···················· ·A·················· ···················· ···················· ···················· ···A················
X 6843 hCoV-19/India/MH-NCCS-NR-8031/2020|EPI_ISL_722181| ···················· ···················· ···················· ···················· ···················· ····················
X 6844 hCoV-19/India/MH-NCCS-NR-6984/2020|EPI_ISL_722182| ···················· ···················· ···················· ···················· ···················· ····················
X 6845 hCoV-19/India/MH-NCCS-NR-7782/2020|EPI_ISL_722183| ···················· ···················· ···················· ···················· ···················· ····················
X 6846 hCoV-19/India/MH-NCCS-NR-5693/2020|EPI_ISL_722184| ···················· ···················· ···················· ···················· ···················· ····················
X 6847 hCoV-19/India/MH-NCCS-NR-7912/2020|EPI_ISL_722185| ···················· ···················· ···················· ···················· ···················· ····················
X 6848 hCoV-19/India/MH-NCCS-NR-7779/2020|EPI_ISL_722186| ···················· ···················· ···················· ···················· ···················· ····················
X 6849 hCoV-19/India/MH-AFCO-7992/2020|EPI_ISL_722187|202 ···················· ···················· ···················· ···················· ···················· ····················
X 6850 hCoV-19/India/MH-AFCO-8223/2020|EPI_ISL_722188|202 ···················· ···················· ···················· ···················· ···················· ····················
X 6851 hCoV-19/India/MH-AFCO-8224/2020|EPI_ISL_722189|202 ···················· ···················· ···················· ···················· ···················· ····················
X 6852 hCoV-19/India/MH-AFCO-8225/2020|EPI_ISL_722190|202 ···················· ···················· ···················· ···················· ···················· ····················
X 6853 hCoV-19/India/MH-AFCO-8699/2020|EPI_ISL_722191|202 ···················· ···················· ···················· ···················· ···················· ····················
X 6854 hCoV-19/India/MH-AFCO-9111/2020|EPI_ISL_722192|202 ···················· ···················· ···················· ···················· ···················· ····················
X 6855 hCoV-19/India/MH-AFCO-9124/2020|EPI_ISL_722193|202 ···················· ···················· ···················· ···················· ···················· ····················
X 6856 hCoV-19/India/MH-AFCO-9126/2020|EPI_ISL_722194|202 ···················· ···················· ···················· ···················· ···················· ····················
X 6857 hCoV-19/India/MH-AFCO-9304/2020|EPI_ISL_722195|202 ···················· ···················· ···················· ···················· ···················· ····················
X 6858 hCoV-19/India/MH-AFCO-9996/2020|EPI_ISL_722196|202 ···················· ···················· ···················· ···················· ···················· ····················
X 6859 hCoV-19/India/MH-AFCO-14763/2020|EPI_ISL_722197|20 ···················· ···················· ···················· ···················· ···················· ····················
X 6860 hCoV-19/India/MH-AFCO-15278/2020|EPI_ISL_722198|20 ···················· ···················· ···················· ···················· ···················· ····················
X 6861 hCoV-19/India/MH-AFCO-15380/2020|EPI_ISL_722199|20 ···················· ···················· ···················· ···················· ···················· ····················
X 6862 hCoV-19/Netherlands/NB-RIVM-20279/2020|EPI_ISL_723 ···················· ···················· ···················· ···················· ···················· ····················
X 6863 hCoV-19/Netherlands/NB-RIVM-20289/2020|EPI_ISL_723 ···················· ···················· ···················· ···················· ···················· ····················
X 6864 hCoV-19/Netherlands/GE-RIVM-20290/2020|EPI_ISL_723 ···················· ···················· ···················· ···················· ···················· ····················
X 6865 hCoV-19/Netherlands/GE-RIVM-20291/2020|EPI_ISL_723 ···················· ···················· ···················· ···················· ···················· ····················
X 6866 hCoV-19/Netherlands/GE-RIVM-20292/2020|EPI_ISL_723 ···················· ···················· ···················· ···················· ···················· ····················
X 6867 hCoV-19/Netherlands/GE-RIVM-20293/2020|EPI_ISL_723 ···················· ···················· ···················· ···················· ···················· ····················
X 6868 hCoV-19/Netherlands/GE-RIVM-20294/2020|EPI_ISL_723 ···················· ···················· ···················· ···················· ···················· ····················
X 6869 hCoV-19/Netherlands/GE-RIVM-20295/2020|EPI_ISL_723 ···················· ···················· ···················· ···················· ···················· ····················
X 6870 hCoV-19/Netherlands/GE-RIVM-20296/2020|EPI_ISL_723 ···················· ···················· ···················· ···················· ···················· ····················
X 6871 hCoV-19/Netherlands/GE-RIVM-20297/2020|EPI_ISL_723 ···················· ···················· ···················· ···················· ···················· ····················
X 6872 hCoV-19/Netherlands/GE-RIVM-20298/2020|EPI_ISL_723 ···················· ···················· ···················· ···················· ···················· ····················
X 6873 hCoV-19/Netherlands/GE-RIVM-20299/2020|EPI_ISL_723 ···················· ···················· ···················· ···················· ···················· ····················
X 6874 hCoV-19/Netherlands/GE-RIVM-20332/2020|EPI_ISL_723 ···················· ···················· ···················· ···················· ···················· ····················
X 6875 hCoV-19/Netherlands/GE-RIVM-20335/2020|EPI_ISL_723 ···················· ···················· ···················· ···················· ···················· ····················
X 6876 hCoV-19/Netherlands/GE-RIVM-20338/2020|EPI_ISL_723 ···················· ···················· ···················· ···················· ···················· ····················
X 6877 hCoV-19/Netherlands/GE-RIVM-20340/2020|EPI_ISL_723 ···················· ···················· ···················· ···················· ···················· ····················
X 6878 hCoV-19/Netherlands/GE-RIVM-20342/2020|EPI_ISL_723 ···················· ···················· ···················· ···················· ···················· ····················
X 6879 hCoV-19/Netherlands/NH-RIVM-20344/2020|EPI_ISL_723 ···················· ···················· ···················· ···················· ···················· ····················
X 6880 hCoV-19/Netherlands/NH-RIVM-20345/2020|EPI_ISL_723 ···················· ···················· ···················· ···················· ···················· ····················
X 6881 hCoV-19/Netherlands/GE-RIVM-20348/2020|EPI_ISL_723 ···················· ···················· ···················· ···················· ···················· ····················
X 6882 hCoV-19/Morocco/LDB-01/2020|EPI_ISL_723469|2020-11 ···················· ···················· ···················· ···················· ···················· ····················
X 6883 hCoV-19/USA/VA-DCLS-2293/2020|EPI_ISL_723470|2020- ···················· ···················· ···················· ···················· ···················· ····················
X 6884 hCoV-19/USA/VA-DCLS-2295/2020|EPI_ISL_723471|2020- ···················· ···················· ···················· ···················· ···················· ····················
X 6885 hCoV-19/USA/VA-DCLS-2296/2020|EPI_ISL_723472|2020- ···················· ···················· ···················· ···················· ···················· ····················
X 6886 hCoV-19/USA/VA-DCLS-2392/2020|EPI_ISL_723473|2020- ···················· ···················· ···················· ···················· ·········T·········· ····················
X 6887 hCoV-19/USA/VA-DCLS-2393/2020|EPI_ISL_723474|2020- ···················· ···················· ···················· ···················· ·········T·········· ····················
X 6888 hCoV-19/USA/VA-DCLS-2394/2020|EPI_ISL_723475|2020- ···················· ···················· ···················· ···················· ·········T·········· ····················
X 6889 hCoV-19/USA/VA-DCLS-2395/2020|EPI_ISL_723476|2020- ···················· ···················· ···················· ···················· ·········T·········· ····················
X 6890 hCoV-19/USA/VA-DCLS-2396/2020|EPI_ISL_723477|2020- ···················· ···················· ···················· ···················· ·········T·········· ····················
X 6891 hCoV-19/USA/VA-DCLS-2397/2020|EPI_ISL_723478|2020- ···················· ···················· ···················· ···················· ·········T·········· ····················
X 6892 hCoV-19/USA/VA-DCLS-2398/2020|EPI_ISL_723479|2020- ···················· ···················· ···················· ···················· ·········T·········· ····················
X 6893 hCoV-19/USA/VA-DCLS-2414/2020|EPI_ISL_723480|2020- ···················· ···················· ···················· ···················· ·········T·········· ····················
X 6894 hCoV-19/USA/VA-DCLS-2415/2020|EPI_ISL_723481|2020- ···················· ···················· ···················· ···················· ·········T·········· ····················
X 6895 hCoV-19/USA/VA-DCLS-2416/2020|EPI_ISL_723482|2020- ···················· ···················· ···················· ···················· ·········T·········· ····················
X 6896 hCoV-19/USA/VA-DCLS-2419/2020|EPI_ISL_723483|2020- ···················· ···················· ···················· ···················· ·········T·········· ····················
X 6897 hCoV-19/USA/VA-DCLS-2420/2020|EPI_ISL_723484|2020- ···················· ···················· ···················· ···················· ·········T·········· ····················
X 6898 hCoV-19/USA/VA-DCLS-2421/2020|EPI_ISL_723485|2020- ···················· ···················· ···················· ···················· ·········T·········· ····················
X 6899 hCoV-19/USA/VA-DCLS-2422/2020|EPI_ISL_723486|2020- ···················· ···················· ···················· ···················· ·········T·········· ····················
X 6900 hCoV-19/USA/VA-DCLS-2423/2020|EPI_ISL_723487|2020- ···················· ···················· ···················· ···················· ·········T·········· ····················
X 6901 hCoV-19/USA/VA-DCLS-2424/2020|EPI_ISL_723488|2020- ···················· ···················· ···················· ···················· ·········T·········· ····················
X 6902 hCoV-19/USA/VA-DCLS-2425/2020|EPI_ISL_723489|2020- ···················· ···················· ···················· ···················· ·········T·········· ····················
X 6903 hCoV-19/USA/VA-DCLS-2431/2020|EPI_ISL_723490|2020- ···················· ···················· ···················· ···················· ·········T·········· ····················
X 6904 hCoV-19/USA/VA-DCLS-2432/2020|EPI_ISL_723491|2020- ···················· ···················· ···················· ···················· ·········T·········· ····················
X 6905 hCoV-19/USA/VA-DCLS-2269/2020|EPI_ISL_723493|2020- ···················· ···················· ···················· ···················· ···················· ····················
X 6906 hCoV-19/USA/VA-DCLS-2191/2020|EPI_ISL_723494|2020- ···················· ···················· ···················· ···················· ···················· ····················
X 6907 hCoV-19/USA/VA-DCLS-2192/2020|EPI_ISL_723495|2020- ···················· ·A·················· ···················· ···················· ···················· ····················
X 6908 hCoV-19/USA/VA-DCLS-2249/2020|EPI_ISL_723496|2020- ···················· ·A·················· ···················· ···················· ···················· ····················
X 6909 hCoV-19/USA/VA-DCLS-2250/2020|EPI_ISL_723497|2020- ···················· ···················· ···················· ···················· ···················· ····················
X 6910 hCoV-19/USA/VA-DCLS-2251/2020|EPI_ISL_723498|2020- ···················· ·A·················· ···················· ···················· ···················· ····················
X 6911 hCoV-19/USA/VA-DCLS-2252/2020|EPI_ISL_723499|2020- ···················· ·A·················· ···················· ···················· ···················· ···A················
X 6912 hCoV-19/USA/VA-DCLS-2290/2020|EPI_ISL_723520|2020- ···················· ···················· ···················· ···················· ···················· ····················
X 6913 hCoV-19/Denmark/DCGC-14012/2020|EPI_ISL_711515|202 ···················· ···················· ···················· ···················· ···················· ····················
```

| | | | | | | | |
|---|---|---|---|---|---|---|---|
| X | 6914 hCoV-19/Denmark/DCGC-14013/2020\|EPI_ISL_711516\|202 | ···················· | ···················· | ····················· | ···················· | ···················· | ···················· |
| X | 6915 hCoV-19/Denmark/DCGC-14014/2020\|EPI_ISL_711517\|202 | ···················· | ···················· | ····················· | ···················· | ···················· | ···················· |
| X | 6916 hCoV-19/Denmark/DCGC-14016/2020\|EPI_ISL_711518\|202 | ···················· | ···················· | ····················· | ···················· | ···················· | ···················· |
| X | 6917 hCoV-19/Denmark/DCGC-14018/2020\|EPI_ISL_711519\|202 | ···················· | ···················· | ····················· | ···················· | ···················· | ···················· |
| X | 6918 hCoV-19/Denmark/DCGC-14019/2020\|EPI_ISL_711520\|202 | ···················· | ···················· | ····················· | ···················· | ···················· | ···················· |
| X | 6919 hCoV-19/Denmark/DCGC-14020/2020\|EPI_ISL_711521\|202 | ···················· | ·A·················· | ····················· | ···················· | ···················· | ···················· |
| X | 6920 hCoV-19/Denmark/DCGC-14021/2020\|EPI_ISL_711522\|202 | ···················· | ···················· | ····················· | ···················· | ···················· | ···················· |
| X | 6921 hCoV-19/Denmark/DCGC-14025/2020\|EPI_ISL_711523\|202 | ···················· | ···················· | ····················· | ···················· | ···················· | ···················· |
| X | 6922 hCoV-19/Denmark/DCGC-14026/2020\|EPI_ISL_711524\|202 | ···················· | ···················· | ····················· | ···················· | ···················· | ···················· |
| X | 6923 hCoV-19/Denmark/DCGC-14027/2020\|EPI_ISL_711525\|202 | ···················· | ···················· | ····················· | ···················· | ···················· | ···················· |
| X | 6924 hCoV-19/Denmark/DCGC-14028/2020\|EPI_ISL_711526\|202 | ···················· | ···················· | ····················· | ···················· | ···················· | ···················· |
| X | 6925 hCoV-19/Denmark/DCGC-14029/2020\|EPI_ISL_711527\|202 | ···················· | ···················· | ····················· | ···················· | ···················· | ···················· |
| X | 6926 hCoV-19/Denmark/DCGC-14030/2020\|EPI_ISL_711528\|202 | ···················· | ···················· | ····················· | ···················· | ···················· | ···················· |
| X | 6927 hCoV-19/Denmark/DCGC-14034/2020\|EPI_ISL_711529\|202 | ···················· | ···················· | ····················· | ···················· | ···················· | ···················· |
| X | 6928 hCoV-19/Denmark/DCGC-14035/2020\|EPI_ISL_711530\|202 | ···················· | ···················· | ····················· | ···················· | ···················· | ···················· |
| X | 6929 hCoV-19/Denmark/DCGC-14040/2020\|EPI_ISL_711531\|202 | ···················· | ···················· | ····················· | ···················· | ···················· | ···················· |
| X | 6930 hCoV-19/Denmark/DCGC-14041/2020\|EPI_ISL_711532\|202 | ···················· | ···················· | ····················· | ···················· | ···················· | ···················· |
| X | 6931 hCoV-19/Denmark/DCGC-14043/2020\|EPI_ISL_711533\|202 | ···················· | ···················· | ····················· | ···················· | ···················· | ···················· |
| X | 6932 hCoV-19/Denmark/DCGC-14044/2020\|EPI_ISL_711534\|202 | ···················· | ···················· | ····················· | ···················· | ···················· | ···················· |
| X | 6933 hCoV-19/Denmark/DCGC-14046/2020\|EPI_ISL_711535\|202 | ···················· | ···················· | ····················· | ···················· | ···················· | ···················· |
| X | 6934 hCoV-19/Denmark/DCGC-14050/2020\|EPI_ISL_711536\|202 | ···················· | ···················· | ····················· | ···················· | ···················· | ···················· |
| X | 6935 hCoV-19/Denmark/DCGC-14058/2020\|EPI_ISL_711540\|202 | ···················· | ···················· | ····················· | ···················· | ···················· | ···················· |
| X | 6936 hCoV-19/Denmark/DCGC-14059/2020\|EPI_ISL_711541\|202 | ···················· | ···················· | ····················· | ···················· | ···················· | ···················· |
| X | 6937 hCoV-19/Denmark/DCGC-14060/2020\|EPI_ISL_711542\|202 | ···················· | ···················· | ····················· | ···················· | ···················· | ···················· |
| X | 6938 hCoV-19/Denmark/DCGC-14061/2020\|EPI_ISL_711543\|202 | ···················· | ···················· | ····················· | ···················· | ···················· | ···················· |
| X | 6939 hCoV-19/Denmark/DCGC-14064/2020\|EPI_ISL_711544\|202 | ···················· | ···················· | ····················· | ···················· | ···················· | ···················· |
| X | 6940 hCoV-19/Denmark/DCGC-14065/2020\|EPI_ISL_711545\|202 | ···················· | ···················· | ····················· | ···················· | ···················· | ···················· |
| X | 6941 hCoV-19/Denmark/DCGC-14069/2020\|EPI_ISL_711548\|202 | ···················· | ···················· | ····················· | ···················· | ···················· | ···················· |
| X | 6942 hCoV-19/Denmark/DCGC-14071/2020\|EPI_ISL_711549\|202 | ···················· | ···················· | ····················· | ···················· | ···················· | ···················· |
| X | 6943 hCoV-19/Denmark/DCGC-14072/2020\|EPI_ISL_711550\|202 | ···················· | ···················· | ····················· | ···················· | ···················· | ···················· |
| X | 6944 hCoV-19/Denmark/DCGC-14074/2020\|EPI_ISL_711551\|202 | ···················· | ···················· | ····················· | ···················· | ···················· | ···················· |
| X | 6945 hCoV-19/Denmark/DCGC-14079/2020\|EPI_ISL_711552\|202 | ···················· | ···················· | ····················· | ···················· | ···················· | ···················· |
| X | 6946 hCoV-19/Denmark/DCGC-14080/2020\|EPI_ISL_711553\|202 | ···················· | ···················· | ····················· | ···················· | ···················· | ···················· |
| X | 6947 hCoV-19/Denmark/DCGC-14081/2020\|EPI_ISL_711554\|202 | ···················· | ···················· | ····················· | ···················· | ···················· | ···················· |
| X | 6948 hCoV-19/Denmark/DCGC-14083/2020\|EPI_ISL_711555\|202 | ···················· | ···················· | ····················· | ···················· | ···················· | ···················· |
| X | 6949 hCoV-19/Denmark/DCGC-14085/2020\|EPI_ISL_711556\|202 | ···················· | ···················· | ····················· | ···················· | ···················· | ···················· |
| X | 6950 hCoV-19/Denmark/DCGC-14087/2020\|EPI_ISL_711557\|202 | ···················· | ···················· | ····················· | ···················· | ···················· | ···················· |
| X | 6951 hCoV-19/Denmark/DCGC-14088/2020\|EPI_ISL_711558\|202 | ···················· | ···················· | ····················· | ···················· | ···················· | ···················· |
| X | 6952 hCoV-19/Denmark/DCGC-14089/2020\|EPI_ISL_711559\|202 | ···················· | ···················· | ····················· | ···················· | ···················· | ···················· |
| X | 6953 hCoV-19/Denmark/DCGC-14090/2020\|EPI_ISL_711560\|202 | ···················· | ···················· | ····················· | ···················· | ···················· | ···················· |
| X | 6954 hCoV-19/Denmark/DCGC-14091/2020\|EPI_ISL_711561\|202 | ···················· | ···················· | ····················· | ···················· | ···················· | ···················· |
| X | 6955 hCoV-19/Denmark/DCGC-14092/2020\|EPI_ISL_711562\|202 | ···················· | ···················· | ····················· | ···················· | ···················· | ···················· |
| X | 6956 hCoV-19/Denmark/DCGC-14093/2020\|EPI_ISL_711563\|202 | ···················· | ···················· | ····················· | ···················· | ···················· | ···················· |
| X | 6957 hCoV-19/Denmark/DCGC-14094/2020\|EPI_ISL_711564\|202 | ···················· | ···················· | ····················· | ···················· | ···················· | ···················· |
| X | 6958 hCoV-19/Denmark/DCGC-14095/2020\|EPI_ISL_711565\|202 | ···················· | ···················· | ····················· | ···················· | ···················· | ···················· |
| X | 6959 hCoV-19/Denmark/DCGC-14097/2020\|EPI_ISL_711566\|202 | ···················· | ···················· | ····················· | ···················· | ···················· | ···················· |
| X | 6960 hCoV-19/Denmark/DCGC-14099/2020\|EPI_ISL_711567\|202 | ···················· | ·A·················· | ····················· | ···················· | ···················· | ···················· |
| X | 6961 hCoV-19/Denmark/DCGC-14101/2020\|EPI_ISL_711568\|202 | ···················· | ···················· | ····················· | ···················· | ···················· | ···················· |
| X | 6962 hCoV-19/Denmark/DCGC-14102/2020\|EPI_ISL_711569\|202 | ···················· | ···················· | ····················· | ···················· | ···················· | ···················· |
| X | 6963 hCoV-19/Denmark/DCGC-14103/2020\|EPI_ISL_711570\|202 | ···················· | ···················· | ····················· | ···················· | ···················· | ···················· |
| X | 6964 hCoV-19/Denmark/DCGC-14104/2020\|EPI_ISL_711571\|202 | ···················· | ···················· | ····················· | ···················· | ···················· | ···················· |
| X | 6965 hCoV-19/Denmark/DCGC-14105/2020\|EPI_ISL_711572\|202 | ···················· | ···················· | ····················· | ···················· | ···················· | ···················· |
| X | 6966 hCoV-19/Denmark/DCGC-14107/2020\|EPI_ISL_711573\|202 | ···················· | ···················· | ····················· | ···················· | ···················· | ···················· |
| X | 6967 hCoV-19/Denmark/DCGC-14109/2020\|EPI_ISL_711574\|202 | ···················· | ···················· | ····················· | ···················· | ···················· | ···················· |
| X | 6968 hCoV-19/Denmark/DCGC-14111/2020\|EPI_ISL_711575\|202 | ···················· | ···················· | ····················· | ···················· | ···················· | ···················· |
| X | 6969 hCoV-19/Denmark/DCGC-14113/2020\|EPI_ISL_711576\|202 | ···················· | ···················· | ····················· | ···················· | ···················· | ···················· |
| X | 6970 hCoV-19/Denmark/DCGC-14114/2020\|EPI_ISL_711577\|202 | ···················· | ···················· | ····················· | ···················· | ···················· | ···················· |
| X | 6971 hCoV-19/Denmark/DCGC-14115/2020\|EPI_ISL_711578\|202 | ···················· | ···················· | ····················· | ···················· | ···················· | ···················· |
| X | 6972 hCoV-19/Denmark/DCGC-14118/2020\|EPI_ISL_711579\|202 | ···················· | ···················· | ····················· | ···················· | ···················· | ···················· |
| X | 6973 hCoV-19/Denmark/DCGC-14119/2020\|EPI_ISL_711580\|202 | ···················· | ···················· | ····················· | ···················· | ···················· | ···················· |
| X | 6974 hCoV-19/Denmark/DCGC-14121/2020\|EPI_ISL_711581\|202 | ···················· | ···················· | ····················· | ···················· | ···················· | ···················· |
| X | 6975 hCoV-19/Denmark/DCGC-14123/2020\|EPI_ISL_711582\|202 | ···················· | ···················· | ····················· | ···················· | ···················· | ···················· |
| X | 6976 hCoV-19/Denmark/DCGC-14125/2020\|EPI_ISL_711583\|202 | ···················· | ···················· | ····················· | ···················· | ···················· | ···················· |
| X | 6977 hCoV-19/Denmark/DCGC-14126/2020\|EPI_ISL_711584\|202 | ···················· | ···················· | ····················· | ···················· | ·····T·············· | ···················· |
| X | 6978 hCoV-19/Denmark/DCGC-14127/2020\|EPI_ISL_711585\|202 | ···················· | ···················· | ····················· | ···················· | ···················· | ···················· |
| X | 6979 hCoV-19/Denmark/DCGC-14129/2020\|EPI_ISL_711586\|202 | ···················· | ···················· | ····················· | ···················· | ···················· | ···················· |
| X | 6980 hCoV-19/Denmark/DCGC-14130/2020\|EPI_ISL_711587\|202 | ···················· | ···················· | ····················· | ···················· | ···················· | ···················· |
| X | 6981 hCoV-19/Denmark/DCGC-14132/2020\|EPI_ISL_711588\|202 | ···················· | ···················· | ····················· | ···················· | ···················· | ···················· |
| X | 6982 hCoV-19/Denmark/DCGC-14138/2020\|EPI_ISL_711589\|202 | ···················· | ·A·················· | ····················· | ···················· | ···················· | ··A················· |
| X | 6983 hCoV-19/Denmark/DCGC-14146/2020\|EPI_ISL_711590\|202 | ···················· | ···················· | ····················· | ···················· | ···················· | ···················· |
| X | 6984 hCoV-19/Denmark/DCGC-14148/2020\|EPI_ISL_711591\|202 | ···················· | ···················· | ····················· | ···················· | ···················· | ···················· |
| X | 6985 hCoV-19/Denmark/DCGC-14150/2020\|EPI_ISL_711592\|202 | ···················· | ···················· | ····················· | ···················· | ···················· | ···················· |
| X | 6986 hCoV-19/Denmark/DCGC-14152/2020\|EPI_ISL_711593\|202 | ···················· | ···················· | ····················· | ···················· | ···················· | ···················· |
| X | 6987 hCoV-19/Denmark/DCGC-14153/2020\|EPI_ISL_711594\|202 | ···················· | ·A·················· | ····················· | ···················· | ···················· | ···················· |
| X | 6988 hCoV-19/Denmark/DCGC-14154/2020\|EPI_ISL_711595\|202 | ···················· | ···················· | ····················· | ···················· | ···················· | ···················· |
| X | 6989 hCoV-19/Denmark/DCGC-14156/2020\|EPI_ISL_711596\|202 | ···················· | ···················· | ····················· | ···················· | ···················· | ···················· |
| X | 6990 hCoV-19/Denmark/DCGC-14157/2020\|EPI_ISL_711597\|202 | ···················· | ···················· | ····················· | ···················· | ···················· | ···················· |
| X | 6991 hCoV-19/Denmark/DCGC-14158/2020\|EPI_ISL_711598\|202 | ···················· | ···················· | ····················· | ···················· | ···················· | ···················· |
| X | 6992 hCoV-19/Denmark/DCGC-14160/2020\|EPI_ISL_711599\|202 | ···················· | ···················· | ····················· | ···················· | ···················· | ···················· |
| X | 6993 hCoV-19/Denmark/DCGC-13511/2020\|EPI_ISL_712846\|202 | ···················· | ···················· | ····················· | ···················· | ···················· | ···················· |

| | | | | | | | |
|---|---|---|---|---|---|---|---|
| X | 6994 hCoV-19/Denmark/DCGC-13512/2020\|EPI_ISL_712847\|202 | ···················· | ···················· | ···················· | ···················· | ···················· | ···················· |
| X | 6995 hCoV-19/Denmark/DCGC-13513/2020\|EPI_ISL_712848\|202 | ···················· | ···················· | ···················· | ···················· | ···················· | ···················· |
| X | 6996 hCoV-19/Denmark/DCGC-13518/2020\|EPI_ISL_712849\|202 | ···················· | ···················· | ···················· | ···················· | ···················· | ···················· |
| X | 6997 hCoV-19/Denmark/DCGC-13519/2020\|EPI_ISL_712850\|202 | ···················· | ···················· | ···················· | ···················· | ···················· | ···················· |
| X | 6998 hCoV-19/Denmark/DCGC-13520/2020\|EPI_ISL_712851\|202 | ···················· | ···················· | ···················· | ···················· | ···················· | ···················· |
| X | 6999 hCoV-19/Denmark/DCGC-13522/2020\|EPI_ISL_712852\|202 | ···················· | ···················· | ···················· | ···················· | ···················· | ···················· |
| X | 7000 hCoV-19/Denmark/DCGC-13523/2020\|EPI_ISL_712853\|202 | ···················· | ···················· | ···················· | ···················· | ···················· | ···················· |
| X | 7001 hCoV-19/Denmark/DCGC-13525/2020\|EPI_ISL_712854\|202 | ···················· | ···················· | ···················· | ···················· | ···················· | ···················· |
| X | 7002 hCoV-19/Denmark/DCGC-13527/2020\|EPI_ISL_712855\|202 | ···················· | ···················· | ···················· | ···················· | ···················· | ···················· |
| X | 7003 hCoV-19/Denmark/DCGC-13528/2020\|EPI_ISL_712856\|202 | ···················· | ···················· | ···················· | ···················· | ···················· | ···················· |
| X | 7004 hCoV-19/Denmark/DCGC-13529/2020\|EPI_ISL_712857\|202 | ···················· | ···················· | ···················· | ···················· | ···················· | ···················· |
| X | 7005 hCoV-19/Denmark/DCGC-13530/2020\|EPI_ISL_712858\|202 | ···················· | ···················· | ···················· | ···················· | ···················· | ···················· |
| X | 7006 hCoV-19/Denmark/DCGC-13532/2020\|EPI_ISL_712859\|202 | ···················· | ···················· | ···················· | ···················· | ···················· | ···················· |
| X | 7007 hCoV-19/Denmark/DCGC-13533/2020\|EPI_ISL_712860\|202 | ···················· | ···················· | ···················· | ···················· | ···················· | ···················· |
| X | 7008 hCoV-19/Denmark/DCGC-13534/2020\|EPI_ISL_712861\|202 | ···················· | ···················· | ···················· | ···················· | ······T············· | ···················· |
| X | 7009 hCoV-19/Denmark/DCGC-13535/2020\|EPI_ISL_712862\|202 | ···················· | ···················· | ···················· | ···················· | ···················· | ···················· |
| X | 7010 hCoV-19/Denmark/DCGC-13536/2020\|EPI_ISL_712863\|202 | ···················· | ···················· | ···················· | ···················· | ···················· | ···················· |
| X | 7011 hCoV-19/Denmark/DCGC-13537/2020\|EPI_ISL_712864\|202 | ···················· | ···················· | ···················· | ···················· | ···················· | ···················· |
| X | 7012 hCoV-19/Denmark/DCGC-13538/2020\|EPI_ISL_712865\|202 | ···················· | ···················· | ···················· | ···················· | ···················· | ···················· |
| X | 7013 hCoV-19/Denmark/DCGC-13539/2020\|EPI_ISL_712866\|202 | ···················· | ···················· | ···················· | ···················· | ···················· | ···················· |
| X | 7014 hCoV-19/Denmark/DCGC-13541/2020\|EPI_ISL_712867\|202 | ···················· | ···················· | ···················· | ···················· | ······T············· | ···················· |
| X | 7015 hCoV-19/Denmark/DCGC-13545/2020\|EPI_ISL_712870\|202 | ···················· | ·A·················· | ···················· | ···················· | ···················· | ···················· |
| X | 7016 hCoV-19/Denmark/DCGC-13549/2020\|EPI_ISL_712871\|202 | ···················· | ···················· | ···················· | ···················· | ···················· | ···················· |
| X | 7017 hCoV-19/Denmark/DCGC-13551/2020\|EPI_ISL_712872\|202 | ···················· | ···················· | ···················· | ···················· | ···················· | ···················· |
| X | 7018 hCoV-19/Denmark/DCGC-13552/2020\|EPI_ISL_712873\|202 | ···················· | ···················· | ···················· | ···················· | ···················· | ···················· |
| X | 7019 hCoV-19/Denmark/DCGC-13553/2020\|EPI_ISL_712874\|202 | ···················· | ···················· | ···················· | ···················· | ···················· | ···················· |
| X | 7020 hCoV-19/Denmark/DCGC-13554/2020\|EPI_ISL_712875\|202 | ···················· | ···················· | ···················· | ···················· | ···················· | ···················· |
| X | 7021 hCoV-19/Denmark/DCGC-13564/2020\|EPI_ISL_712879\|202 | ···················· | ···················· | ···················· | ···················· | ···················· | ···················· |
| X | 7022 hCoV-19/Denmark/DCGC-13568/2020\|EPI_ISL_712880\|202 | ···················· | ···················· | ···················· | ···················· | ···················· | ···················· |
| X | 7023 hCoV-19/Denmark/DCGC-13569/2020\|EPI_ISL_712881\|202 | ···················· | ···················· | ···················· | ···················· | ···················· | ···················· |
| X | 7024 hCoV-19/Denmark/DCGC-13570/2020\|EPI_ISL_712882\|202 | ···················· | ···················· | ···················· | ···················· | ···················· | ···················· |
| X | 7025 hCoV-19/Denmark/DCGC-13572/2020\|EPI_ISL_712883\|202 | ···················· | ···················· | ···················· | ···················· | ···················· | ···················· |
| X | 7026 hCoV-19/Denmark/DCGC-13573/2020\|EPI_ISL_712884\|202 | ···················· | ···················· | ···················· | ···················· | ···················· | ···················· |
| X | 7027 hCoV-19/Denmark/DCGC-13574/2020\|EPI_ISL_712885\|202 | ···················· | ···················· | ···················· | ···················· | ···················· | ···················· |
| X | 7028 hCoV-19/Denmark/DCGC-13575/2020\|EPI_ISL_712886\|202 | ···················· | ···················· | ···················· | ···················· | ···················· | ···················· |
| X | 7029 hCoV-19/Denmark/DCGC-13576/2020\|EPI_ISL_712887\|202 | ···················· | ···················· | ···················· | ···················· | ···················· | ···················· |
| X | 7030 hCoV-19/Denmark/DCGC-13577/2020\|EPI_ISL_712888\|202 | ···················· | ···················· | ···················· | ···················· | ···················· | ···················· |
| X | 7031 hCoV-19/Denmark/DCGC-13578/2020\|EPI_ISL_712889\|202 | ···················· | ···················· | ···················· | ···················· | ···················· | ···················· |
| X | 7032 hCoV-19/Denmark/DCGC-13580/2020\|EPI_ISL_712890\|202 | ···················· | ···················· | ···················· | ···················· | ···················· | ···················· |
| X | 7033 hCoV-19/Denmark/DCGC-13581/2020\|EPI_ISL_712891\|202 | ···················· | ···················· | ···················· | ···················· | ···················· | ···················· |
| X | 7034 hCoV-19/Denmark/DCGC-13584/2020\|EPI_ISL_712892\|202 | ···················· | ···················· | ···················· | ···················· | ···················· | ···················· |
| X | 7035 hCoV-19/Denmark/DCGC-13585/2020\|EPI_ISL_712893\|202 | ···················· | ···················· | ···················· | ···················· | ···················· | ···················· |
| X | 7036 hCoV-19/Denmark/DCGC-13586/2020\|EPI_ISL_712894\|202 | ···················· | ·A·················· | ···················· | ···················· | ···················· | ···················· |
| X | 7037 hCoV-19/Denmark/DCGC-13589/2020\|EPI_ISL_712895\|202 | ···················· | ···················· | ···················· | ···················· | ···················· | ···················· |
| X | 7038 hCoV-19/Denmark/DCGC-13592/2020\|EPI_ISL_712896\|202 | ···················· | ···················· | ···················· | ···················· | ···················· | ···················· |
| X | 7039 hCoV-19/Denmark/DCGC-13594/2020\|EPI_ISL_712897\|202 | ···················· | ···················· | ···················· | ···················· | ···················· | ···················· |
| X | 7040 hCoV-19/Denmark/DCGC-13595/2020\|EPI_ISL_712898\|202 | ···················· | ···················· | ···················· | ···················· | ···················· | ···················· |
| X | 7041 hCoV-19/Denmark/DCGC-13597/2020\|EPI_ISL_712899\|202 | ···················· | ···················· | ···················· | ···················· | ···················· | ···················· |
| X | 7042 hCoV-19/Australia/NSW3628/2020\|EPI_ISL_717701\|2020 | ···················· | ···················· | ···················· | ···················· | ···················· | ···················· |
| X | 7043 hCoV-19/Australia/NSW3629/2020\|EPI_ISL_717702\|2020 | ···················· | ·········A·········· | ···················· | ···················· | ···················· | ···················· |
| X | 7044 hCoV-19/Australia/NSW3631/2020\|EPI_ISL_717703\|2020 | ···················· | ·A·················· | ···················· | ···················· | ···················· | ···A················ |
| X | 7045 hCoV-19/Canada/ON-SLB3109/2020\|EPI_ISL_717727\|2020 | ···················· | ···················· | ···················· | ···················· | ···················· | ···················· |
| X | 7046 hCoV-19/Canada/ON-SLB3079/2020\|EPI_ISL_717728\|2020 | ···················· | ···················· | ···················· | ···················· | ···················· | ···················· |
| X | 7047 hCoV-19/Canada/ON-SLB3096/2020\|EPI_ISL_717729\|2020 | ···················· | ···················· | ···················· | ···················· | ···················· | ···················· |
| X | 7048 hCoV-19/Canada/ON-SLB3125/2020\|EPI_ISL_717730\|2020 | ···················· | ···················· | ···················· | ···················· | ···················· | ···················· |
| X | 7049 hCoV-19/Canada/ON-SLB3167/2020\|EPI_ISL_717731\|2020 | ···················· | ···················· | ···················· | ···················· | ···················· | ···················· |
| X | 7050 hCoV-19/Canada/ON-SLB3153/2020\|EPI_ISL_717732\|2020 | ···················· | ···················· | ···················· | ···················· | ···················· | ···················· |
| X | 7051 hCoV-19/Canada/ON-SLB3139/2020\|EPI_ISL_717733\|2020 | ···················· | ···················· | ···················· | ···················· | ···················· | ···················· |
| X | 7052 hCoV-19/Canada/ON-SLB3242/2020\|EPI_ISL_717734\|2020 | ···················· | ·A·················· | ···················· | ···················· | ···················· | ···A················ |
| X | 7053 hCoV-19/Canada/ON-SLB3101/2020\|EPI_ISL_717735\|2020 | ···················· | ···················· | ···················· | ···················· | ···················· | ···················· |
| X | 7054 hCoV-19/Canada/ON-SLB3119/2020\|EPI_ISL_717736\|2020 | ···················· | ···················· | ···················· | ···················· | ···················· | ···················· |
| X | 7055 hCoV-19/Canada/ON-SLB3243/2020\|EPI_ISL_717737\|2020 | ···················· | ·A·················· | ···················· | ···················· | ···················· | ···A················ |
| X | 7056 hCoV-19/Canada/ON-SLB3140/2020\|EPI_ISL_717738\|2020 | ···················· | ···················· | ···················· | ···················· | ···················· | ···················· |
| X | 7057 hCoV-19/Canada/ON-SLB3144/2020\|EPI_ISL_717739\|2020 | ···················· | ···················· | ···················· | ···················· | ···················· | ···················· |
| X | 7058 hCoV-19/India/MH-AFCO-15534/2020\|EPI_ISL_722200\|20 | ···················· | ···················· | ···················· | ···················· | ···················· | ···················· |
| X | 7059 hCoV-19/Bosnia and Herzegovina/AGC-MOS-002/2020\|EP | ···················· | ···················· | ···················· | ···················· | ···················· | ···················· |
| X | 7060 hCoV-19/Switzerland/ZH-UZH-IMV100/2020\|EPI_ISL_722 | ···················· | ···················· | ···················· | ···················· | ···················· | ···················· |
| X | 7061 hCoV-19/Switzerland/LU-UZH-IMV101/2020\|EPI_ISL_722 | ···················· | ···················· | ···················· | ···················· | ···················· | ···················· |
| X | 7062 hCoV-19/Switzerland/ZH-UZH-IMV102/2020\|EPI_ISL_722 | ···················· | ···················· | ···················· | ···················· | ···················· | ···················· |
| X | 7063 hCoV-19/Switzerland/SZ-UZH-IMV103/2020\|EPI_ISL_722 | ···················· | ···················· | ···················· | ···················· | ···················· | ···················· |
| X | 7064 hCoV-19/Switzerland/AG-UZH-IMV104/2020\|EPI_ISL_722 | ···················· | ···················· | ···················· | ···················· | ···················· | ···················· |
| X | 7065 hCoV-19/Switzerland/ZH-UZH-IMV105/2020\|EPI_ISL_722 | ···················· | ···················· | ···················· | ···················· | ···················· | ···················· |
| X | 7066 hCoV-19/Switzerland/ZH-UZH-IMV106/2020\|EPI_ISL_722 | ···················· | ···················· | ···················· | ···················· | ···················· | ···················· |
| X | 7067 hCoV-19/Bosnia and Herzegovina/AGC-SAR-002/2020\|EP | ···················· | ···················· | ···················· | ···················· | ···················· | ···················· |
| X | 7068 hCoV-19/Spain/AR-IBV-98010617/2020\|EPI_ISL_722224\| | ···················· | ···················· | ···················· | ···················· | ···················· | ···················· |
| X | 7069 hCoV-19/Spain/AR-IBV-98010623/2020\|EPI_ISL_722225\| | ···················· | ···················· | ···················· | ···················· | ···················· | ···················· |
| X | 7070 hCoV-19/Spain/AR-IBV-98010625/2020\|EPI_ISL_722226\| | ···················· | ···················· | ···················· | ···················· | ···················· | ···················· |
| X | 7071 hCoV-19/Spain/AR-IBV-98010626/2020\|EPI_ISL_722227\| | ···················· | ···················· | ···················· | ···················· | ···················· | ···················· |
| X | 7072 hCoV-19/Spain/AR-IBV-98010592/2020\|EPI_ISL_722228\| | ···················· | ···················· | ···················· | ···················· | ···················· | ···················· |
| X | 7073 hCoV-19/Spain/AR-IBV-98010593/2020\|EPI_ISL_722229\| | ···················· | ···················· | ···················· | ···················· | ···················· | ···················· |

X 7074 hCoV-19/Spain/AR-IBV-98010561/2020|EPI_ISL_722230| ................... ................... ................... ................... ................... ...................
X 7075 hCoV-19/Spain/AR-IBV-98010578/2020|EPI_ISL_722231| ................... ................... ................... ................... ................... ...................
X 7076 hCoV-19/Spain/AR-IBV-98010641/2020|EPI_ISL_722232| ................... ................... ................... ................... ................... ...................
X 7077 hCoV-19/Spain/AS-IBV-98006619/2020|EPI_ISL_722241| ................... ................... ................... ................... ................... ...................
X 7078 hCoV-19/Spain/AR-IBV-98010577/2020|EPI_ISL_722242| ................... ................... ................... ................... ................... ...................
X 7079 hCoV-19/Spain/AR-IBV-98010579/2020|EPI_ISL_722243| ................... ................... ................... ................... ................... ...................
X 7080 hCoV-19/Spain/AR-IBV-98010580/2020|EPI_ISL_722244| ................... ................... ................... ................... ................... ...................
X 7081 hCoV-19/Spain/AR-IBV-98010582/2020|EPI_ISL_722245| ................... ................... ................... ................... ................... ...................
X 7082 hCoV-19/Spain/AR-IBV-98010585/2020|EPI_ISL_722246| ................... ................... ................... ................... ................... ...................
X 7083 hCoV-19/Spain/AR-IBV-98010589/2020|EPI_ISL_722247| ................... A.................. ................... ................... ................... ...................
X 7084 hCoV-19/Spain/AR-IBV-98010590/2020|EPI_ISL_722248| ................... ................... ................... ................... ................... ...................
X 7085 hCoV-19/Spain/AR-IBV-98010594/2020|EPI_ISL_722249| ................... ................... ................... ................... ................... ...................
X 7086 hCoV-19/Spain/AR-IBV-98010596/2020|EPI_ISL_722250| ................... ................... ................... ................... ................... ...................
X 7087 hCoV-19/Spain/AR-IBV-98010599/2020|EPI_ISL_722251| ................... ................... ................... ................... ................... ...................
X 7088 hCoV-19/Spain/AR-IBV-98010603/2020|EPI_ISL_722252| ................... ................... ................... ................... ................... ...................
X 7089 hCoV-19/Spain/AR-IBV-98010604/2020|EPI_ISL_722253| ................... ................... ................... ................... ................... ...................
X 7090 hCoV-19/Spain/AR-IBV-98010608/2020|EPI_ISL_722254| ................... ................... ................... ................... ................... ...................
X 7091 hCoV-19/Spain/AR-IBV-98010613/2020|EPI_ISL_722255| ................... A.................. ................... ................... ................... ...................
X 7092 hCoV-19/Spain/AR-IBV-98010614/2020|EPI_ISL_722256| ................... ................... ................... ................... ................... ...................
X 7093 hCoV-19/Spain/AR-IBV-98010615/2020|EPI_ISL_722257| ................... ................... ................... ................... ................... ...................
X 7094 hCoV-19/Spain/AR-IBV-98010619/2020|EPI_ISL_722258| ................... ................... ................... ................... ................... ...................
X 7095 hCoV-19/Spain/AR-IBV-98010620/2020|EPI_ISL_722259| ................... ................... ................... ................... ................... ...................
X 7096 hCoV-19/Spain/AR-IBV-98010621/2020|EPI_ISL_722260| ................... ................... ................... ................... ................... ...................
X 7097 hCoV-19/Spain/AR-IBV-98010622/2020|EPI_ISL_722261| ................... ................... ................... ................... ................... ...................
X 7098 hCoV-19/Spain/AR-IBV-98010624/2020|EPI_ISL_722262| ................... ................... ................... ................... ................... ...................
X 7099 hCoV-19/Spain/AR-IBV-98010634/2020|EPI_ISL_722263| ................... ................... ................... ................... ................... ...................
X 7100 hCoV-19/Spain/AR-IBV-98010640/2020|EPI_ISL_722264| ................... ................... ................... ................... ................... ...................
X 7101 hCoV-19/Spain/AR-IBV-98010651/2020|EPI_ISL_722265| ................... ................... ................... ................... ................... ...................
X 7102 hCoV-19/Spain/AR-IBV-98010652/2020|EPI_ISL_722266| ................... ................... ................... ................... ................... ...................
X 7104 hCoV-19/Spain/AR-IBV-98010673/2020|EPI_ISL_722268| ................... ................... ................... ................... ................... ...................
X 7105 hCoV-19/Spain/AR-IBV-98010683/2020|EPI_ISL_722269| ................... ................... ................... ................... ................... ...................
X 7106 hCoV-19/Spain/AR-IBV-98010715/2020|EPI_ISL_722270| ................... ................... ................... ................... ................... ...................
X 7107 hCoV-19/Spain/AR-IBV-98010736/2020|EPI_ISL_722271| ................... ................... ................... ................... ................... ...................
X 7109 hCoV-19/USA/VA-DCLS-2253/2020|EPI_ISL_723500|2020- ................... .A................. ................... ................... ................... ..A................
X 7110 hCoV-19/USA/VA-DCLS-2254/2020|EPI_ISL_723501|2020- ................... .A................. ................... ................... ................... ...................
X 7111 hCoV-19/USA/VA-DCLS-2255/2020|EPI_ISL_723502|2020- ................... ................... ................... ................... ................... ...................
X 7112 hCoV-19/USA/VA-DCLS-2256/2020|EPI_ISL_723503|2020- ................... ................... ................... ................... ................... ...................
X 7113 hCoV-19/USA/VA-DCLS-2257/2020|EPI_ISL_723504|2020- ................... .A................. ................... ................... ................... ...................
X 7114 hCoV-19/USA/VA-DCLS-2258/2020|EPI_ISL_723505|2020- ................... .A................. ................... ................... ................... ..A................
X 7115 hCoV-19/USA/VA-DCLS-2259/2020|EPI_ISL_723506|2020- ................... ................... ................... ................... ................... ...................
X 7116 hCoV-19/USA/VA-DCLS-2263/2020|EPI_ISL_723507|2020- ................... .A................. ................... ................... ................... ...................
X 7117 hCoV-19/USA/VA-DCLS-2264/2020|EPI_ISL_723508|2020- ................... ................... ................... ................... ................... ...................
X 7118 hCoV-19/USA/VA-DCLS-2265/2020|EPI_ISL_723509|2020- ................... ................... ................... ................... ................... ...................
X 7119 hCoV-19/USA/VA-DCLS-2266/2020|EPI_ISL_723510|2020- .................A. ................... ................... ................... ................... ................... 1
X 7120 hCoV-19/USA/VA-DCLS-2267/2020|EPI_ISL_723511|2020- ................... ................... ................... ................... ................... ...................
X 7121 hCoV-19/USA/VA-DCLS-2271/2020|EPI_ISL_723512|2020- ................... ................... ................... ................... ................... ...................
X 7122 hCoV-19/USA/VA-DCLS-2272/2020|EPI_ISL_723513|2020- ................... .A................. ................... ................... ................... ...................
X 7123 hCoV-19/USA/VA-DCLS-2273/2020|EPI_ISL_723514|2020- ................... ................... ................... ................... ................... ...................
X 7124 hCoV-19/USA/VA-DCLS-2276/2020|EPI_ISL_723515|2020- ................... ................... ................... ................... ................... ...................
X 7125 hCoV-19/USA/VA-DCLS-2277/2020|EPI_ISL_723516|2020- ................... .A................. ................... ................... ................... ...................
X 7126 hCoV-19/USA/VA-DCLS-2281/2020|EPI_ISL_723517|2020- ................... ................... ................... ................... ................... ...................
X 7127 hCoV-19/USA/VA-DCLS-2285/2020|EPI_ISL_723518|2020- ................... ................... ................... ................... ................... ...................
X 7128 hCoV-19/USA/VA-DCLS-2287/2020|EPI_ISL_723519|2020- ................... .A................. ................... ................... ................... ...................
X 7129 hCoV-19/England/ALDP-B1CB05/2020|EPI_ISL_723530|20 ................... ................... ................... ................... .....T............. ...................
X 7130 hCoV-19/England/ALDP-B1D041/2020|EPI_ISL_723531|20 ................... .A................. ................... ................... ................... ..A................
X 7131 hCoV-19/Wales/ALDP-B1DCE3/2020|EPI_ISL_723535|2020 ................... ................... ................... ................... ................... ...................
X 7132 hCoV-19/Wales/ALDP-B1DCF2/2020|EPI_ISL_723536|2020 ................... ................... ................... ................... ................... ...................
X 7133 hCoV-19/Wales/ALDP-B1DD3B/2020|EPI_ISL_723540|2020 ................... ................... ................... ................... ................... ...................
X 7134 hCoV-19/Wales/ALDP-B1DDA4/2020|EPI_ISL_723546|2020 ................... ................... ................... ................... ................... ...................
X 7135 hCoV-19/England/ALDP-B1DF62/2020|EPI_ISL_723551|20 ................... ................... ................... ................... ................... ...................
X 7136 hCoV-19/Wales/ALDP-B1DF71/2020|EPI_ISL_723552|2020 ................... ................... ................... ................... ................... ...................
X 7137 hCoV-19/Wales/ALDP-B1DF9F/2020|EPI_ISL_723554|2020 ................... ................... ................... ................... ................... ...................
X 7138 hCoV-19/Wales/ALDP-B1DFBD/2020|EPI_ISL_723555|2020 ................... ................... ................... ................... ................... ...................
X 7139 hCoV-19/England/ALDP-B1DFDB/2020|EPI_ISL_723557|20 ................... ................... ................... ................... ................... ...................
X 7140 hCoV-19/England/ALDP-B5D91D/2020|EPI_ISL_723558|20 ................... ................... ................... ................... ................... ...................
X 7141 hCoV-19/England/ALDP-B5D995/2020|EPI_ISL_723559|20 ................... ................... ................... ................... ................... ...................
X 7142 hCoV-19/England/ALDP-B5DB35/2020|EPI_ISL_723561|20 ................... ................... ................... ................... ................... ...................
X 7143 hCoV-19/England/ALDP-B5DB44/2020|EPI_ISL_723562|20 ................... ................... ................... ................... ................... ...................
X 7144 hCoV-19/England/ALDP-B5DB53/2020|EPI_ISL_723563|20 ................... ................... ................... ................... ................... ...................
X 7145 hCoV-19/England/ALDP-B5DB80/2020|EPI_ISL_723565|20 ................... ................... ................... ................... ................... ...................
X 7146 hCoV-19/England/ALDP-B5DB9F/2020|EPI_ISL_723566|20 ................... ................... ................... ................... ................... ...................
X 7147 hCoV-19/England/ALDP-B5DBAE/2020|EPI_ISL_723567|20 ................... ................... ................... ................... ................... ...................
X 7148 hCoV-19/England/ALDP-B5DBBD/2020|EPI_ISL_723568|20 ................... ................... ................... ................... ................... ...................
X 7149 hCoV-19/England/ALDP-B5DBCC/2020|EPI_ISL_723569|20 ................... ................... ................... ................... ................... ...................
X 7150 hCoV-19/England/ALDP-B5DBEA/2020|EPI_ISL_723571|20 ................... ................... ................... ................... ................... ...................
X 7151 hCoV-19/England/ALDP-B5DBF9/2020|EPI_ISL_723572|20 ................... ................... ................... ................... ................... ...................
X 7152 hCoV-19/England/ALDP-B5DC14/2020|EPI_ISL_723574|20 ................... ................... ................... ................... ................... ...................
X 7153 hCoV-19/England/ALDP-B5DC32/2020|EPI_ISL_723576|20 ................... ................... ................... ................... ................... ...................
X 7154 hCoV-19/England/ALDP-B5DC6F/2020|EPI_ISL_723578|20 ................... .A................. ................... ................... ................... ..A................
X 7155 hCoV-19/England/ALDP-B5DC8D/2020|EPI_ISL_723579|20 ................... ................... ................... ................... ................... ...................

| | | | | | | | | |
|---|---|---|---|---|---|---|---|---|
| X | 7156 | hCoV-19/England/ALDP-B5DCD8/2020\|EPI_ISL_723581\|20 | .................... | .................... | .................... | .................... | .................... | .................... |
| X | 7157 | hCoV-19/England/ALDP-B5DCE7/2020\|EPI_ISL_723582\|20 | .................... | .................... | .................... | .................... | .................... | .................... |
| X | 7158 | hCoV-19/England/ALDP-B5DCF6/2020\|EPI_ISL_723583\|20 | .................... | .................... | .................... | .................... | .................... | .................... |
| X | 7159 | hCoV-19/England/ALDP-B5DD11/2020\|EPI_ISL_723584\|20 | .................... | .................... | .................... | .................... | .................... | .................... |
| X | 7160 | hCoV-19/England/ALDP-B5DD20/2020\|EPI_ISL_723585\|20 | .................... | .................... | .................... | .................... | .................... | .................... |
| X | 7161 | hCoV-19/England/ALDP-B5DD3F/2020\|EPI_ISL_723586\|20 | .................... | .................... | .................... | .................... | .................... | .................... |
| X | 7162 | hCoV-19/England/ALDP-B5DD4E/2020\|EPI_ISL_723587\|20 | .................... | .................... | .................... | .................... | .................... | .................... |
| X | 7163 | hCoV-19/England/ALDP-B5DD6C/2020\|EPI_ISL_723589\|20 | .................... | .A.................. | .................... | .................... | .................... | ..A................. |
| X | 7164 | hCoV-19/England/ALDP-B5DD7B/2020\|EPI_ISL_723590\|20 | .................... | .................... | .................... | .................... | .................... | .................... |
| X | 7165 | hCoV-19/England/ALDP-B5DDB7/2020\|EPI_ISL_723593\|20 | .................... | .................... | .................... | .................... | .................... | .................... |
| X | 7166 | hCoV-19/England/ALDP-B5DDD5/2020\|EPI_ISL_723594\|20 | .................... | .................... | .................... | .................... | .................... | .................... |
| X | 7167 | hCoV-19/England/ALDP-B5DE0F/2020\|EPI_ISL_723596\|20 | .................... | .................... | .................... | .................... | .................... | .................... |
| X | 7168 | hCoV-19/England/ALDP-B5DE1E/2020\|EPI_ISL_723597\|20 | .................... | .................... | .................... | .................... | .................... | .................... |
| X | 7169 | hCoV-19/Denmark/DCGC-14161/2020\|EPI_ISL_711600\|202 | .................... | .................... | .................... | .................... | .................... | .................... |
| X | 7170 | hCoV-19/Denmark/DCGC-14163/2020\|EPI_ISL_711601\|202 | .................... | .................... | .................... | .................... | .................... | .................... |
| X | 7171 | hCoV-19/Denmark/DCGC-14164/2020\|EPI_ISL_711602\|202 | .................... | .................... | .................... | .................... | .................... | .................... |
| X | 7172 | hCoV-19/Denmark/DCGC-14169/2020\|EPI_ISL_711603\|202 | .................... | .................... | .................... | .................... | .................... | .................... |
| X | 7173 | hCoV-19/Denmark/DCGC-14171/2020\|EPI_ISL_711604\|202 | .................... | .................... | .................... | .................... | .................... | .................... |
| X | 7174 | hCoV-19/Denmark/DCGC-14174/2020\|EPI_ISL_711605\|202 | .................... | .................... | .................... | .................... | .................... | .................... |
| X | 7175 | hCoV-19/Denmark/DCGC-14176/2020\|EPI_ISL_711606\|202 | .................... | .................... | .................... | .................... | .................... | .................... |
| X | 7176 | hCoV-19/Denmark/DCGC-14178/2020\|EPI_ISL_711607\|202 | .................... | .................... | .................... | .................... | .................... | .................... |
| X | 7177 | hCoV-19/Denmark/DCGC-14179/2020\|EPI_ISL_711608\|202 | .................... | .................... | .................... | .................... | .................... | .................... |
| X | 7178 | hCoV-19/Denmark/DCGC-14180/2020\|EPI_ISL_711609\|202 | .................... | .................... | .................... | .................... | .................... | .................... |
| X | 7179 | hCoV-19/Denmark/DCGC-14181/2020\|EPI_ISL_711610\|202 | .................... | .................... | .................... | .................... | .................... | .................... |
| X | 7180 | hCoV-19/Denmark/DCGC-14182/2020\|EPI_ISL_711611\|202 | .................... | .................... | .................... | .................... | .................... | .................... |
| X | 7181 | hCoV-19/Denmark/DCGC-14183/2020\|EPI_ISL_711612\|202 | .................... | .................... | .................... | .................... | .................... | .................... |
| X | 7182 | hCoV-19/Denmark/DCGC-14186/2020\|EPI_ISL_711613\|202 | .................... | .................... | .................... | .................... | .................... | .................... |
| X | 7183 | hCoV-19/Denmark/DCGC-14189/2020\|EPI_ISL_711614\|202 | .................... | .A.................. | .................... | .................... | .................... | ..A................. |
| X | 7184 | hCoV-19/Denmark/DCGC-14192/2020\|EPI_ISL_711615\|202 | .................... | .................... | .................... | .................... | .................... | .................... |
| X | 7185 | hCoV-19/Denmark/DCGC-14193/2020\|EPI_ISL_711616\|202 | .................... | .A.................. | .................... | .................... | .................... | .................... |
| X | 7186 | hCoV-19/Denmark/DCGC-14194/2020\|EPI_ISL_711617\|202 | .................... | .................... | .................... | .................... | .................... | .................... |
| X | 7187 | hCoV-19/Denmark/DCGC-14195/2020\|EPI_ISL_711618\|202 | .................... | .................... | .................... | .................... | .................... | .................... |
| X | 7188 | hCoV-19/Denmark/DCGC-14197/2020\|EPI_ISL_711619\|202 | .................... | .................... | .................... | .................... | .................... | .................... |
| X | 7189 | hCoV-19/Denmark/DCGC-14199/2020\|EPI_ISL_711620\|202 | .................... | .................... | .................... | .................... | .................... | .................... |
| X | 7190 | hCoV-19/Denmark/DCGC-14200/2020\|EPI_ISL_711621\|202 | .................... | .................... | .................... | .................... | .................... | .................... |
| X | 7191 | hCoV-19/Denmark/DCGC-14201/2020\|EPI_ISL_711622\|202 | .................... | .................... | .................... | .................... | .................... | .................... |
| X | 7192 | hCoV-19/Denmark/DCGC-14202/2020\|EPI_ISL_711623\|202 | .................... | .................... | .................... | .................... | .................... | .................... |
| X | 7193 | hCoV-19/Denmark/DCGC-14203/2020\|EPI_ISL_711624\|202 | .................... | .................... | .................... | .................... | .................... | .................... |
| X | 7194 | hCoV-19/Denmark/DCGC-14205/2020\|EPI_ISL_711625\|202 | .................... | .................... | .................... | .................... | .................... | .................... |
| X | 7195 | hCoV-19/Denmark/DCGC-14209/2020\|EPI_ISL_711626\|202 | .................... | .................... | .................... | .................... | .................... | .................... |
| X | 7196 | hCoV-19/Denmark/DCGC-14212/2020\|EPI_ISL_711627\|202 | .................... | .................... | .................... | .................... | .................... | .................... |
| X | 7197 | hCoV-19/Denmark/DCGC-14213/2020\|EPI_ISL_711628\|202 | .................... | .................... | .................... | .................... | .................... | .................... |
| X | 7198 | hCoV-19/Denmark/DCGC-14214/2020\|EPI_ISL_711629\|202 | .................... | .................... | .................... | .................... | .................... | .................... |
| X | 7199 | hCoV-19/Denmark/DCGC-14216/2020\|EPI_ISL_711630\|202 | .................... | .................... | .................... | .................... | .................... | .................... |
| X | 7200 | hCoV-19/Denmark/DCGC-14217/2020\|EPI_ISL_711631\|202 | .................... | .................... | .................... | .................... | .................... | .................... |
| X | 7201 | hCoV-19/Denmark/DCGC-14218/2020\|EPI_ISL_711632\|202 | .................... | .................... | .................... | .................... | .................... | .................... |
| X | 7202 | hCoV-19/Denmark/DCGC-14219/2020\|EPI_ISL_711633\|202 | .................... | .................... | .................... | .................... | .................... | .................... |
| X | 7203 | hCoV-19/Denmark/DCGC-14220/2020\|EPI_ISL_711634\|202 | .................... | .................... | .................... | .................... | .................... | .................... |
| X | 7204 | hCoV-19/Denmark/DCGC-14222/2020\|EPI_ISL_711635\|202 | .................... | .................... | .................... | .................... | .................... | .................... |
| X | 7205 | hCoV-19/Denmark/DCGC-14224/2020\|EPI_ISL_711636\|202 | .................... | .................... | .................... | .................... | .................... | .................... |
| X | 7206 | hCoV-19/Denmark/DCGC-14225/2020\|EPI_ISL_711637\|202 | .................... | .................... | .................... | .................... | .................... | .................... |
| X | 7207 | hCoV-19/Denmark/DCGC-14226/2020\|EPI_ISL_711638\|202 | .................... | .................... | .................... | .................... | .................... | .................... |
| X | 7208 | hCoV-19/Denmark/DCGC-14227/2020\|EPI_ISL_711639\|202 | .................... | .................... | .................... | .................... | .................... | .................... |
| X | 7209 | hCoV-19/Denmark/DCGC-14228/2020\|EPI_ISL_711640\|202 | .................... | .................... | .................... | .................... | .................... | .................... |
| X | 7210 | hCoV-19/Denmark/DCGC-14229/2020\|EPI_ISL_711641\|202 | .................... | .................... | .................... | .................... | .................... | .................... |
| X | 7211 | hCoV-19/Denmark/DCGC-14230/2020\|EPI_ISL_711642\|202 | .................... | .................... | .................... | .................... | .................... | .................... |
| X | 7212 | hCoV-19/Denmark/DCGC-14231/2020\|EPI_ISL_711643\|202 | .................... | .A.................. | .................... | .................... | .................... | ..A................. |
| X | 7213 | hCoV-19/Denmark/DCGC-14233/2020\|EPI_ISL_711644\|202 | .................... | .................... | .................... | .................... | .................... | .................... |
| X | 7214 | hCoV-19/Denmark/DCGC-14235/2020\|EPI_ISL_711645\|202 | .................... | .................... | .................... | .................... | .................... | .................... |
| X | 7215 | hCoV-19/Denmark/DCGC-14236/2020\|EPI_ISL_711646\|202 | .................... | .................... | .................... | .................... | .................... | .................... |
| X | 7216 | hCoV-19/Denmark/DCGC-14238/2020\|EPI_ISL_711647\|202 | .................... | .................... | .................... | .................... | .................... | .................... |
| X | 7217 | hCoV-19/Denmark/DCGC-14243/2020\|EPI_ISL_711649\|202 | .................... | .................... | .................... | .................... | .................... | .................... |
| X | 7218 | hCoV-19/Denmark/DCGC-14255/2020\|EPI_ISL_711658\|202 | .................... | .................... | .................... | .................... | .................... | .................... |
| X | 7219 | hCoV-19/Denmark/DCGC-14257/2020\|EPI_ISL_711659\|202 | .................... | .................... | .................... | .................... | .................... | .................... |
| X | 7220 | hCoV-19/Denmark/DCGC-14260/2020\|EPI_ISL_711660\|202 | .................... | .................... | .................... | .................... | .................... | .................... |
| X | 7221 | hCoV-19/Denmark/DCGC-14263/2020\|EPI_ISL_711661\|202 | .................... | ...A................ | .................... | .................... | .................... | .................... |
| X | 7222 | hCoV-19/Denmark/DCGC-14264/2020\|EPI_ISL_711662\|202 | .................... | .................... | .................... | .................... | .................... | .................... |
| X | 7223 | hCoV-19/Denmark/DCGC-14265/2020\|EPI_ISL_711663\|202 | .................... | .A.................. | .................... | .................... | .................... | .................... |
| X | 7224 | hCoV-19/Denmark/DCGC-14267/2020\|EPI_ISL_711664\|202 | .................... | .A.................. | .................... | .................... | .................... | ..A................. |
| X | 7225 | hCoV-19/Denmark/DCGC-14271/2020\|EPI_ISL_711665\|202 | .................... | .................... | .................... | .................... | .................... | .................... |
| X | 7226 | hCoV-19/Denmark/DCGC-14273/2020\|EPI_ISL_711666\|202 | .................... | .................... | .................... | .................... | .................... | .................... |
| X | 7227 | hCoV-19/Denmark/DCGC-14275/2020\|EPI_ISL_711667\|202 | .................... | .................... | .................... | .................... | .................... | .................... |
| X | 7228 | hCoV-19/Denmark/DCGC-14276/2020\|EPI_ISL_711668\|202 | .................... | .................... | .................... | .................... | .................... | .................... |
| X | 7229 | hCoV-19/Denmark/DCGC-14279/2020\|EPI_ISL_711669\|202 | .................... | .................... | .................... | .................... | .................... | .................... |
| X | 7230 | hCoV-19/Denmark/DCGC-14280/2020\|EPI_ISL_711670\|202 | .................... | .................... | .................... | .................... | .................... | .................... |
| X | 7231 | hCoV-19/Denmark/DCGC-14281/2020\|EPI_ISL_711671\|202 | .................... | .................... | .................... | .................... | .................... | .................... |
| X | 7232 | hCoV-19/Denmark/DCGC-14282/2020\|EPI_ISL_711672\|202 | .................... | .................... | .................... | .................... | .................... | .................... |
| X | 7233 | hCoV-19/Denmark/DCGC-14285/2020\|EPI_ISL_711673\|202 | .................... | .................... | .................... | .................... | .................... | .................... |
| X | 7234 | hCoV-19/Denmark/DCGC-14290/2020\|EPI_ISL_711674\|202 | .................... | .................... | .................... | .................... | .................... | .................... |
| X | 7235 | hCoV-19/Denmark/DCGC-14291/2020\|EPI_ISL_711675\|202 | .................... | .................... | .................... | .................... | .................... | .................... |

| | | | | | | | | |
|---|---|---|---|---|---|---|---|---|
| X | 7236 | hCoV-19/Denmark/DCGC-14295/2020\|EPI_ISL_711676\|202 | ···················· | ···················· | ···················· | ···················· | ···················· | ···················· |
| X | 7237 | hCoV-19/Denmark/DCGC-14296/2020\|EPI_ISL_711677\|202 | ···················· | ···················· | ···················· | ···················· | ···················· | ···················· |
| X | 7238 | hCoV-19/Denmark/DCGC-14297/2020\|EPI_ISL_711678\|202 | ···················· | ···················· | ···················· | ···················· | ···················· | ···················· |
| X | 7239 | hCoV-19/Denmark/DCGC-14298/2020\|EPI_ISL_711679\|202 | ···················· | ···················· | ···················· | ···················· | ···················· | ···················· |
| X | 7240 | hCoV-19/Denmark/DCGC-13598/2020\|EPI_ISL_712900\|202 | ···················· | ···················· | ···················· | ···················· | ···················· | ···················· |
| X | 7241 | hCoV-19/Denmark/DCGC-13599/2020\|EPI_ISL_712901\|202 | ···················· | ···················· | ···················· | ···················· | ···················· | ···················· |
| X | 7242 | hCoV-19/Denmark/DCGC-13600/2020\|EPI_ISL_712902\|202 | ···················· | ···················· | ···················· | ···················· | ···················· | ···················· |
| X | 7243 | hCoV-19/Denmark/DCGC-13602/2020\|EPI_ISL_712903\|202 | ···················· | ···················· | ···················· | ···················· | ···················· | ···················· |
| X | 7244 | hCoV-19/Denmark/DCGC-13604/2020\|EPI_ISL_712904\|202 | ···················· | ···················· | ···················· | ···················· | ···················· | ···················· |
| X | 7245 | hCoV-19/Denmark/DCGC-13605/2020\|EPI_ISL_712905\|202 | ···················· | ···················· | ···················· | ···················· | ···················· | ···················· |
| X | 7246 | hCoV-19/Denmark/DCGC-13606/2020\|EPI_ISL_712906\|202 | ···················· | ···················· | ···················· | ···················· | ···················· | ···················· |
| X | 7247 | hCoV-19/Denmark/DCGC-13607/2020\|EPI_ISL_712907\|202 | ···················· | ···················· | ···················· | ···················· | ···················· | ···················· |
| X | 7248 | hCoV-19/Denmark/DCGC-13608/2020\|EPI_ISL_712908\|202 | ···················· | ···················· | ···················· | ···················· | ···················· | ···················· |
| X | 7249 | hCoV-19/Denmark/DCGC-13610/2020\|EPI_ISL_712909\|202 | ···················· | ···················· | ···················· | ···················· | ···················· | ···················· |
| X | 7250 | hCoV-19/Denmark/DCGC-13611/2020\|EPI_ISL_712910\|202 | ···················· | ···················· | ···················· | ···················· | ···················· | ···················· |
| X | 7251 | hCoV-19/Denmark/DCGC-13612/2020\|EPI_ISL_712911\|202 | ···················· | ···················· | ···················· | ···················· | ···················· | ···················· |
| X | 7252 | hCoV-19/Denmark/DCGC-13615/2020\|EPI_ISL_712912\|202 | ···················· | ···················· | ···················· | ···················· | ···················· | ···················· |
| X | 7253 | hCoV-19/Denmark/DCGC-13616/2020\|EPI_ISL_712913\|202 | ···················· | ···················· | ···················· | ···················· | ···················· | ···················· |
| X | 7254 | hCoV-19/Denmark/DCGC-13617/2020\|EPI_ISL_712914\|202 | ···················· | ···················· | ···················· | ···················· | ···················· | ···················· |
| X | 7255 | hCoV-19/Denmark/DCGC-13618/2020\|EPI_ISL_712915\|202 | ···················· | ···················· | ···················· | ···················· | ···················· | ···················· |
| X | 7256 | hCoV-19/Denmark/DCGC-13619/2020\|EPI_ISL_712916\|202 | ···················· | ···················· | ···················· | ···················· | ···················· | ···················· |
| X | 7257 | hCoV-19/Denmark/DCGC-13620/2020\|EPI_ISL_712917\|202 | ···················· | ···················· | ···················· | ···················· | ···················· | ···················· |
| X | 7258 | hCoV-19/Denmark/DCGC-13623/2020\|EPI_ISL_712918\|202 | ···················· | ···················· | ···················· | ···················· | ···················· | ···················· |
| X | 7259 | hCoV-19/Denmark/DCGC-13624/2020\|EPI_ISL_712919\|202 | ···················· | ···················· | ···················· | ···················· | ···················· | ···················· |
| X | 7260 | hCoV-19/Denmark/DCGC-13625/2020\|EPI_ISL_712920\|202 | ···················· | ·A·················· | ···················· | ···················· | ···················· | ···················· |
| X | 7261 | hCoV-19/Denmark/DCGC-13626/2020\|EPI_ISL_712921\|202 | ···················· | ···················· | ···················· | ···················· | ···················· | ···················· |
| X | 7262 | hCoV-19/Denmark/DCGC-13627/2020\|EPI_ISL_712922\|202 | ···················· | ···················· | ···················· | ···················· | ···················· | ···················· |
| X | 7263 | hCoV-19/Denmark/DCGC-13628/2020\|EPI_ISL_712923\|202 | ···················· | ···················· | ···················· | ···················· | ···················· | ···················· |
| X | 7264 | hCoV-19/Denmark/DCGC-13629/2020\|EPI_ISL_712924\|202 | ···················· | ···················· | ···················· | ···················· | ···················· | ···················· |
| X | 7265 | hCoV-19/Denmark/DCGC-13631/2020\|EPI_ISL_712925\|202 | ···················· | ···················· | ···················· | ···················· | ···················· | ···················· |
| X | 7266 | hCoV-19/Denmark/DCGC-13632/2020\|EPI_ISL_712926\|202 | ···················· | ···················· | ···················· | ···················· | ···················· | ···················· |
| X | 7267 | hCoV-19/Denmark/DCGC-13633/2020\|EPI_ISL_712927\|202 | ···················· | ···················· | ···················· | ···················· | ···················· | ···················· |
| X | 7268 | hCoV-19/Denmark/DCGC-13634/2020\|EPI_ISL_712928\|202 | ···················· | ···················· | ···················· | ···················· | ···················· | ···················· |
| X | 7269 | hCoV-19/Denmark/DCGC-13637/2020\|EPI_ISL_712929\|202 | ···················· | ·A·················· | ···················· | ···················· | ···················· | ···················· |
| X | 7270 | hCoV-19/Denmark/DCGC-13642/2020\|EPI_ISL_712930\|202 | ···················· | ···················· | ···················· | ···················· | ···················· | ···················· |
| X | 7271 | hCoV-19/Denmark/DCGC-13644/2020\|EPI_ISL_712931\|202 | ···················· | ···················· | ···················· | ···················· | ···················· | ···················· |
| X | 7272 | hCoV-19/Denmark/DCGC-13645/2020\|EPI_ISL_712932\|202 | ···················· | ···················· | ···················· | ···················· | ···················· | ···················· |
| X | 7273 | hCoV-19/Denmark/DCGC-13646/2020\|EPI_ISL_712933\|202 | ···················· | ···················· | ···················· | ···················· | ···················· | ···················· |
| X | 7274 | hCoV-19/Denmark/DCGC-13648/2020\|EPI_ISL_712934\|202 | ···················· | ···················· | ···················· | ···················· | ···················· | ···················· |
| X | 7275 | hCoV-19/Denmark/DCGC-13649/2020\|EPI_ISL_712935\|202 | ···················· | ···················· | ···················· | ···················· | ···················· | ···················· |
| X | 7276 | hCoV-19/Denmark/DCGC-13650/2020\|EPI_ISL_712936\|202 | ···················· | ···················· | ···················· | ···················· | ···················· | ···················· |
| X | 7277 | hCoV-19/Denmark/DCGC-13651/2020\|EPI_ISL_712937\|202 | ···················· | ···················· | ···················· | ···················· | ···················· | ···················· |
| X | 7278 | hCoV-19/Denmark/DCGC-13652/2020\|EPI_ISL_712938\|202 | ···················· | ···················· | ···················· | ···················· | ···················· | ···················· |
| X | 7279 | hCoV-19/Denmark/DCGC-13653/2020\|EPI_ISL_712939\|202 | ···················· | ···················· | ···················· | ···················· | ···················· | ···················· |
| X | 7280 | hCoV-19/Denmark/DCGC-13654/2020\|EPI_ISL_712940\|202 | ···················· | ···················· | ···················· | ···················· | ···················· | ···················· |
| X | 7281 | hCoV-19/Denmark/DCGC-13655/2020\|EPI_ISL_712941\|202 | ···················· | ···················· | ···················· | ···················· | ···················· | ···················· |
| X | 7282 | hCoV-19/Denmark/DCGC-13656/2020\|EPI_ISL_712942\|202 | ···················· | ···················· | ···················· | ···················· | ···················· | ···················· |
| X | 7283 | hCoV-19/Denmark/DCGC-13659/2020\|EPI_ISL_712943\|202 | ···················· | ···················· | ···················· | ···················· | ···················· | ···················· |
| X | 7284 | hCoV-19/Denmark/DCGC-13660/2020\|EPI_ISL_712944\|202 | ···················· | ···················· | ···················· | ···················· | ···················· | ···················· |
| X | 7285 | hCoV-19/Denmark/DCGC-13666/2020\|EPI_ISL_712945\|202 | ···················· | ···················· | ···················· | ···················· | ···················· | ···················· |
| X | 7286 | hCoV-19/Denmark/DCGC-13667/2020\|EPI_ISL_712946\|202 | ···················· | ···················· | ··T················· | ···················· | ···················· | ···················· |
| X | 7287 | hCoV-19/Denmark/DCGC-13669/2020\|EPI_ISL_712947\|202 | ···················· | ···················· | ···················· | ···················· | ···················· | ···················· |
| X | 7288 | hCoV-19/Denmark/DCGC-13671/2020\|EPI_ISL_712948\|202 | ···················· | ···················· | ···················· | ···················· | ···················· | ···················· |
| X | 7289 | hCoV-19/Denmark/DCGC-13672/2020\|EPI_ISL_712949\|202 | ···················· | ···················· | ···················· | ···················· | ···················· | ···················· |
| X | 7290 | hCoV-19/Denmark/DCGC-13674/2020\|EPI_ISL_712950\|202 | ···················· | ···················· | ···················· | ···················· | ···················· | ···················· |
| X | 7291 | hCoV-19/Denmark/DCGC-13675/2020\|EPI_ISL_712951\|202 | ···················· | ···················· | ···················· | ···················· | ···················· | ···················· |
| X | 7292 | hCoV-19/Denmark/DCGC-13676/2020\|EPI_ISL_712952\|202 | ···················· | ···················· | ···················· | ···················· | ···················· | ···················· |
| X | 7293 | hCoV-19/Denmark/DCGC-13677/2020\|EPI_ISL_712953\|202 | ···················· | ···················· | ···················· | ···················· | ···················· | ···················· |
| X | 7294 | hCoV-19/Denmark/DCGC-13678/2020\|EPI_ISL_712954\|202 | ···················· | ···················· | ···················· | ···················· | ···················· | ···················· |
| X | 7295 | hCoV-19/Denmark/DCGC-13679/2020\|EPI_ISL_712955\|202 | ···················· | ···················· | ···················· | ···················· | ···················· | ···················· |
| X | 7296 | hCoV-19/Denmark/DCGC-13680/2020\|EPI_ISL_712956\|202 | ···················· | ···················· | ···················· | ···················· | ···················· | ···················· |
| X | 7297 | hCoV-19/Denmark/DCGC-13681/2020\|EPI_ISL_712957\|202 | ···················· | ···················· | ···················· | ···················· | ···················· | ···················· |
| X | 7298 | hCoV-19/Denmark/DCGC-13683/2020\|EPI_ISL_712958\|202 | ···················· | ···················· | ···················· | ···················· | ···················· | ···················· |
| X | 7299 | hCoV-19/Denmark/DCGC-13685/2020\|EPI_ISL_712959\|202 | ···················· | ···················· | ···················· | ···················· | ···················· | ···················· |
| X | 7300 | hCoV-19/Denmark/DCGC-13686/2020\|EPI_ISL_712960\|202 | ···················· | ···················· | ···················· | ···················· | ···················· | ···················· |
| X | 7301 | hCoV-19/Denmark/DCGC-13689/2020\|EPI_ISL_712961\|202 | ···················· | ···················· | ···················· | ···················· | ···················· | ···················· |
| X | 7302 | hCoV-19/Denmark/DCGC-13691/2020\|EPI_ISL_712962\|202 | ···················· | ···················· | ···················· | ···················· | ···················· | ···················· |
| X | 7303 | hCoV-19/Denmark/DCGC-13692/2020\|EPI_ISL_712963\|202 | ···················· | ···················· | ···················· | ···················· | ···················· | ···················· |
| X | 7304 | hCoV-19/Denmark/DCGC-13695/2020\|EPI_ISL_712964\|202 | ···················· | ···················· | ···················· | ···················· | ···················· | ···················· |
| X | 7305 | hCoV-19/Denmark/DCGC-13696/2020\|EPI_ISL_712965\|202 | ···················· | ···················· | ···················· | ···················· | ···················· | ···················· |
| X | 7306 | hCoV-19/Denmark/DCGC-13699/2020\|EPI_ISL_712966\|202 | ···················· | ···················· | ···················· | ···················· | ···················· | ···················· |
| X | 7307 | hCoV-19/Denmark/DCGC-13700/2020\|EPI_ISL_712967\|202 | ···················· | ···················· | ···················· | ···················· | ···················· | ···················· |
| X | 7308 | hCoV-19/Denmark/DCGC-13703/2020\|EPI_ISL_712968\|202 | ···················· | ···················· | ···················· | ···················· | ···················· | ···················· |
| X | 7309 | hCoV-19/Denmark/DCGC-13704/2020\|EPI_ISL_712969\|202 | ···················· | ···················· | ···················· | ···················· | ···················· | ···················· |
| X | 7310 | hCoV-19/Denmark/DCGC-13705/2020\|EPI_ISL_712970\|202 | ···················· | ···················· | ···················· | ···················· | ···················· | ···················· |
| X | 7311 | hCoV-19/Denmark/DCGC-13706/2020\|EPI_ISL_712971\|202 | ···················· | ···················· | ···················· | ···················· | ···················· | ···················· |
| X | 7312 | hCoV-19/Denmark/DCGC-13707/2020\|EPI_ISL_712972\|202 | ···················· | ···················· | ···················· | ···················· | ···················· | ···················· |
| X | 7313 | hCoV-19/Denmark/DCGC-13708/2020\|EPI_ISL_712973\|202 | ···················· | ···················· | ···················· | ···················· | ···················· | ···················· |
| X | 7314 | hCoV-19/Denmark/DCGC-13711/2020\|EPI_ISL_712974\|202 | ···················· | ···················· | ···················· | ···················· | ···················· | ···················· |
| X | 7315 | hCoV-19/Denmark/DCGC-13712/2020\|EPI_ISL_712975\|202 | ···················· | ···················· | ···················· | ···················· | ···················· | ···················· |

| | | | | | | | |
|---|---|---|---|---|---|---|---|
| X | 7316 hCoV-19/Denmark/DCGC-13713/2020\|EPI_ISL_712976\|202 | .................... | .................... | ..................... | .................... | .................... | .................... |
| X | 7317 hCoV-19/Denmark/DCGC-13714/2020\|EPI_ISL_712977\|202 | .................... | .................... | ..................... | .................... | .................... | .................... |
| X | 7318 hCoV-19/Denmark/DCGC-13716/2020\|EPI_ISL_712978\|202 | .................... | .A.................. | ..................... | .................... | .................... | ...A................ |
| X | 7319 hCoV-19/Denmark/DCGC-13717/2020\|EPI_ISL_712979\|202 | .................... | .................... | ..................... | .................... | .................... | .................... |
| X | 7320 hCoV-19/Denmark/DCGC-13735/2020\|EPI_ISL_712989\|202 | .................... | .................... | ..................... | .................... | .................... | .................... |
| X | 7321 hCoV-19/Denmark/DCGC-13736/2020\|EPI_ISL_712990\|202 | .................... | .................... | ..................... | .................... | .................... | .................... |
| X | 7322 hCoV-19/Denmark/DCGC-13737/2020\|EPI_ISL_712991\|202 | .................... | .................... | ..................... | .................... | .................... | .................... |
| X | 7323 hCoV-19/Denmark/DCGC-13739/2020\|EPI_ISL_712992\|202 | .................... | .................... | ..................... | .................... | .................... | .................... |
| X | 7324 hCoV-19/Denmark/DCGC-13740/2020\|EPI_ISL_712993\|202 | .................... | .................... | ..................... | .................... | .................... | .................... |
| X | 7325 hCoV-19/Denmark/DCGC-13741/2020\|EPI_ISL_712994\|202 | .................... | .................... | ..................... | .................... | .................... | .................... |
| X | 7326 hCoV-19/Denmark/DCGC-13745/2020\|EPI_ISL_712995\|202 | .................... | .................... | ..................... | .................... | .................... | .................... |
| X | 7327 hCoV-19/Denmark/DCGC-13746/2020\|EPI_ISL_712996\|202 | .................... | .................... | ..................... | .................... | .................... | .................... |
| X | 7328 hCoV-19/Denmark/DCGC-13748/2020\|EPI_ISL_712997\|202 | .................... | .................... | ..................... | .................... | .................... | .................... |
| X | 7329 hCoV-19/Denmark/DCGC-13749/2020\|EPI_ISL_712998\|202 | .................... | .................... | ..................... | .................... | .................... | .................... |
| X | 7330 hCoV-19/Denmark/DCGC-13750/2020\|EPI_ISL_712999\|202 | .................... | .................... | ..................... | .................... | .................... | .................... |
| X | 7331 hCoV-19/Australia/NSW3634/2020\|EPI_ISL_717704\|2020 | .................... | .................... | ..................... | .................... | .................... | .................... |
| X | 7332 hCoV-19/Australia/NSW3638/2020\|EPI_ISL_717705\|2020 | .................... | .................... | ..................... | .................... | .................... | .................... |
| X | 7333 hCoV-19/Australia/NSW3639/2020\|EPI_ISL_717706\|2020 | .................... | .................... | ..................... | .................... | .................... | .................... |
| X | 7334 hCoV-19/Australia/NSW3642/2020\|EPI_ISL_717707\|2020 | .................... | .A.................. | ..................... | .................... | .................... | ...A................ |
| X | 7335 hCoV-19/Australia/NSW3644/2020\|EPI_ISL_717708\|2020 | .................... | .................... | ..................... | .................... | .................... | .................... |
| X | 7336 hCoV-19/Australia/NSW3646/2020\|EPI_ISL_717710\|2020 | .................... | .................... | ..................... | .................... | .................... | .................... |
| X | 7337 hCoV-19/Australia/NSW3648/2020\|EPI_ISL_717712\|2020 | .................... | .A.................. | ..................... | .................... | .................... | ...A................ |
| X | 7338 hCoV-19/Australia/NSW3664/2020\|EPI_ISL_717713\|2020 | .................... | .A.................. | ..................... | .................... | .................... | ...A................ |
| X | 7339 hCoV-19/mink/Canada/A/2020\|EPI_ISL_717714\|2020-12- | .................... | .................... | ..................... | .................... | .................... | .................... |
| X | 7340 hCoV-19/mink/Canada/B/2020\|EPI_ISL_717715\|2020-12- | .................... | .................... | ..................... | .................... | .................... | .................... |
| X | 7341 hCoV-19/mink/Canada/C/2020\|EPI_ISL_717716\|2020-12- | .................... | .................... | ..................... | .................... | .................... | .................... |
| X | 7342 hCoV-19/mink/Canada/D/2020\|EPI_ISL_717717\|2020-12- | .................... | .................... | ..................... | .................... | .................... | .................... |
| X | 7343 hCoV-19/Canada/ON-SLB3099/2020\|EPI_ISL_717740\|2020 | .................... | .................... | ..................... | .................... | .................... | .................... |
| X | 7344 hCoV-19/Canada/ON-SLB3244/2020\|EPI_ISL_717741\|2020 | .................... | .................... | ..................... | .................... | .................... | .................... |
| X | 7345 hCoV-19/Canada/ON-SLB3105/2020\|EPI_ISL_717742\|2020 | .................... | .................... | ..................... | .................... | .................... | .................... |
| X | 7346 hCoV-19/Canada/ON-SLB3132/2020\|EPI_ISL_717743\|2020 | .................... | .................... | ..................... | .................... | .................... | .................... |
| X | 7347 hCoV-19/Canada/ON-SLB3168/2020\|EPI_ISL_717744\|2020 | .................... | .................... | ..................... | .................... | .................... | .................... |
| X | 7348 hCoV-19/Canada/ON-SLB3103/2020\|EPI_ISL_717745\|2020 | .................... | .................... | ..................... | .................... | .................... | .................... |
| X | 7349 hCoV-19/Canada/ON-SLB3076/2020\|EPI_ISL_717746\|2020 | .................... | .................... | ..................... | .................... | .................... | .................... |
| X | 7350 hCoV-19/Canada/ON-SLB3095/2020\|EPI_ISL_717747\|2020 | .................... | .................... | ..................... | .................... | .................... | .................... |
| X | 7351 hCoV-19/Canada/ON-SLB3131/2020\|EPI_ISL_717748\|2020 | .................... | .................... | ..................... | .................... | .................... | .................... |
| X | 7352 hCoV-19/Canada/ON-SLB3081/2020\|EPI_ISL_717749\|2020 | .................... | .................... | ..................... | .................... | .................... | .................... |
| X | 7353 hCoV-19/Canada/ON-SLB3086/2020\|EPI_ISL_717750\|2020 | .................... | .................... | ..................... | .................... | .................... | .................... |
| X | 7354 hCoV-19/Canada/ON-SLB3090/2020\|EPI_ISL_717751\|2020 | .................... | .................... | ..................... | .................... | .................... | .................... |
| X | 7355 hCoV-19/Canada/ON-SLB3245/2020\|EPI_ISL_717752\|2020 | .................... | .A.................. | ..................... | .................... | .................... | ...A................ |
| X | 7356 hCoV-19/Canada/ON-SLB3124/2020\|EPI_ISL_717753\|2020 | .................... | .................... | ..................... | .................... | .................... | .................... |
| X | 7357 hCoV-19/Canada/ON-SLB3104/2020\|EPI_ISL_717754\|2020 | .................... | .................... | ..................... | .................... | .................... | .................... |
| X | 7358 hCoV-19/Canada/ON-SLB3114/2020\|EPI_ISL_717755\|2020 | .................... | .................... | ..................... | .................... | .................... | .................... |
| X | 7359 hCoV-19/Canada/ON-SLB3098/2020\|EPI_ISL_717756\|2020 | .................... | .................... | ..................... | .................... | .................... | .................... |
| X | 7360 hCoV-19/Canada/ON-SLB3110/2020\|EPI_ISL_717757\|2020 | .................... | .................... | ..................... | .................... | .................... | .................... |
| X | 7361 hCoV-19/Canada/ON-SLB3141/2020\|EPI_ISL_717758\|2020 | .................... | .................... | ..................... | .................... | .................... | .................... |
| X | 7362 hCoV-19/Canada/ON-SLB3106/2020\|EPI_ISL_717759\|2020 | .................... | .................... | ..................... | .................... | .................... | .................... |
| X | 7363 hCoV-19/Canada/ON-SLB3082/2020\|EPI_ISL_717760\|2020 | .................... | .................... | ..................... | .................... | .................... | .................... |
| X | 7364 hCoV-19/Canada/ON-SLB3147/2020\|EPI_ISL_717761\|2020 | .................... | .................... | ..................... | .................... | .................... | .................... |
| X | 7365 hCoV-19/USA/WA-UW-15204/2020\|EPI_ISL_717766\|2020-0 | .................... | .................... | ..................... | .................... | .................... | .................... |
| X | 7366 hCoV-19/USA/WA-UW-18499/2020\|EPI_ISL_717767\|2020-0 | .................... | .................... | ..................... | .................... | .................... | .................... |
| X | 7367 hCoV-19/USA/WA-UW-2044/2020\|EPI_ISL_717768\|2020-03 | .................... | .................... | ..................... | .................... | .................... | .................... |
| X | 7368 hCoV-19/USA/WA-UW-23215/2020\|EPI_ISL_717769\|2020-0 | .................... | .A.................. | ..................... | .................... | .................... | .................... |
| X | 7369 hCoV-19/USA/WA-UW-27118/2020\|EPI_ISL_717770\|2020-0 | .................... | .A.................. | ..................... | .................... | .................... | .................... |
| X | 7370 hCoV-19/USA/WA-UW-27490/2020\|EPI_ISL_717771\|2020-0 | .................... | .................... | ..................... | .................... | .................... | .................... |
| X | 7371 hCoV-19/USA/WA-UW-27494/2020\|EPI_ISL_717772\|2020-0 | .................... | .A.................. | ..................... | .................... | .................... | .................... |
| X | 7372 hCoV-19/USA/WA-UW-28259/2020\|EPI_ISL_717773\|2020-0 | .................... | .A.................. | ..................... | .................... | .................... | .................... |
| X | 7373 hCoV-19/USA/WA-UW-28263/2020\|EPI_ISL_717774\|2020-0 | .................... | .A.................. | ..................... | .................... | .................... | .................... |
| X | 7374 hCoV-19/USA/WA-UW-28264/2020\|EPI_ISL_717775\|2020-0 | .................... | .A.................. | ..................... | .................... | .................... | .................... |
| X | 7375 hCoV-19/USA/WA-UW-29719/2020\|EPI_ISL_717777\|2020-0 | .................... | .................... | ..................... | .................... | .................... | .................... |
| X | 7376 hCoV-19/USA/WA-UW-29741/2020\|EPI_ISL_717778\|2020-0 | .................... | .A.................. | ..................... | .................... | .................... | .................... |
| X | 7377 hCoV-19/USA/WA-UW-29758/2020\|EPI_ISL_717780\|2020-0 | .................... | .................... | ..................... | .................... | .................... | .................... |
| X | 7378 hCoV-19/USA/WA-UW-29772/2020\|EPI_ISL_717781\|2020-0 | .................... | .................... | ..................... | .................... | .................... | .................... |
| X | 7379 hCoV-19/USA/WA-UW-37930/2020\|EPI_ISL_717782\|2020-1 | .................... | .A.................. | ..................... | .................... | .................... | .................... |
| X | 7380 hCoV-19/USA/WA-UW-757/2020\|EPI_ISL_717783\|2020-03- | .................... | .................... | ..................... | .................... | .................... | .................... |
| X | 7381 hCoV-19/USA/WA-UW-9774/2020\|EPI_ISL_717784\|2020-05 | .................... | .................... | ..................... | .................... | .................... | .................... |
| X | 7382 hCoV-19/England/ALDP-B5DE4B/2020\|EPI_ISL_723600\|20 | .................... | .................... | ..................... | .................... | .................... | .................... |
| X | 7383 hCoV-19/England/ALDP-B5DE69/2020\|EPI_ISL_723601\|20 | .................... | .................... | ..................... | .................... | .................... | .................... |
| X | 7384 hCoV-19/England/ALDP-B5DE87/2020\|EPI_ISL_723603\|20 | .................... | .................... | ..................... | .................... | ......T............. | .................... |
| X | 7385 hCoV-19/England/ALDP-B5DE96/2020\|EPI_ISL_723604\|20 | .................... | .................... | ..................... | .................... | .................... | .................... |
| X | 7386 hCoV-19/England/ALDP-B5DEA5/2020\|EPI_ISL_723605\|20 | .................... | .................... | ..................... | .................... | .................... | .................... |
| X | 7387 hCoV-19/England/ALDP-B5DEC3/2020\|EPI_ISL_723606\|20 | .................... | .........T.......... | ..................... | .................... | .................... | .................... |
| X | 7388 hCoV-19/England/ALDP-B5DED2/2020\|EPI_ISL_723607\|20 | .................... | .................... | ..................... | .................... | .................... | .................... |
| X | 7389 hCoV-19/England/ALDP-B5DF0C/2020\|EPI_ISL_723610\|20 | .................... | .................... | ..................... | .................... | .................... | .................... |
| X | 7390 hCoV-19/England/ALDP-B5DF2A/2020\|EPI_ISL_723612\|20 | .................... | .................... | ..................... | .................... | .................... | .................... |
| X | 7391 hCoV-19/England/ALDP-B5DF57/2020\|EPI_ISL_723615\|20 | .................... | .................... | ..................... | .................... | .................... | .................... |
| X | 7392 hCoV-19/England/ALDP-B5DF93/2020\|EPI_ISL_723618\|20 | .................... | .................... | ..................... | .................... | .................... | .................... |
| X | 7393 hCoV-19/England/ALDP-B5DFA2/2020\|EPI_ISL_723619\|20 | .................... | .A.................. | ..................... | .................... | .................... | ...A................ |
| X | 7394 hCoV-19/England/ALDP-B5DFB1/2020\|EPI_ISL_723620\|20 | .................... | .................... | ..................... | .................... | .................... | .................... |
| X | 7395 hCoV-19/England/ALDP-B5DFEE/2020\|EPI_ISL_723621\|20 | .................... | .................... | ..................... | .................... | .................... | .................... |

| | | | | | | | | |
|---|---|---|---|---|---|---|---|---|
| X | 7396 | hCoV-19/England/ALDP-B5DFFD/2020\|EPI_ISL_723622\|20 | ···················· | ···················· | ···················· | ···················· | ···················· | ···················· |
| X | 7397 | hCoV-19/England/ALDP-B5E017/2020\|EPI_ISL_723624\|20 | ···················· | ···················· | ···················· | ···················· | ···················· | ···················· |
| X | 7398 | hCoV-19/England/ALDP-B5E0AE/2020\|EPI_ISL_723630\|20 | ···················· | ···················· | ···················· | ···················· | ···················· | ···················· |
| X | 7399 | hCoV-19/England/ALDP-B5E0CC/2020\|EPI_ISL_723631\|20 | ···················· | ···················· | ···················· | ···················· | ···················· | ···················· |
| X | 7400 | hCoV-19/England/ALDP-B5E0EA/2020\|EPI_ISL_723632\|20 | ···················· | ···················· | ···················· | ···················· | ···················· | ···················· |
| X | 7401 | hCoV-19/England/ALDP-B5E0F9/2020\|EPI_ISL_723633\|20 | ···················· | ···················· | ···················· | ···················· | ···················· | ···················· |
| X | 7402 | hCoV-19/England/ALDP-B5E123/2020\|EPI_ISL_723634\|20 | ···················· | ···················· | ···················· | ···················· | ···················· | ···················· |
| X | 7403 | hCoV-19/England/ALDP-B5E132/2020\|EPI_ISL_723635\|20 | ···················· | ···················· | ···················· | ···················· | ···················· | ···················· |
| X | 7404 | hCoV-19/England/ALDP-B5E17E/2020\|EPI_ISL_723636\|20 | ···················· | ···················· | ···················· | ···················· | ···················· | ···················· |
| X | 7405 | hCoV-19/England/ALDP-B5E19C/2020\|EPI_ISL_723637\|20 | ···················· | ···················· | ···················· | ···················· | ···················· | ···················· |
| X | 7406 | hCoV-19/England/ALDP-B5E1BA/2020\|EPI_ISL_723638\|20 | ···················· | ···················· | ···················· | ···················· | ···················· | ···················· |
| X | 7407 | hCoV-19/England/ALDP-B5E1C9/2020\|EPI_ISL_723639\|20 | ···················· | ···················· | ···················· | ···················· | ···················· | ···················· |
| X | 7408 | hCoV-19/England/ALDP-B5E1D8/2020\|EPI_ISL_723640\|20 | ···················· | ···················· | ···················· | ···················· | ···················· | ···················· |
| X | 7409 | hCoV-19/England/ALDP-B5E1E7/2020\|EPI_ISL_723641\|20 | ···················· | ···················· | ···················· | ···················· | ···················· | ···················· |
| X | 7410 | hCoV-19/England/ALDP-B5E220/2020\|EPI_ISL_723642\|20 | ···················· | ···················· | ···················· | ···················· | ···················· | ···················· |
| X | 7411 | hCoV-19/England/ALDP-B5E23F/2020\|EPI_ISL_723643\|20 | ···················· | ···················· | ···················· | ···················· | ···················· | ···················· |
| X | 7412 | hCoV-19/England/ALDP-B5E24E/2020\|EPI_ISL_723644\|20 | ···················· | ···················· | ···················· | ···················· | ···················· | ···················· |
| X | 7413 | hCoV-19/England/ALDP-B5E26C/2020\|EPI_ISL_723645\|20 | ···················· | ···················· | ···················· | ···················· | ···················· | ···················· |
| X | 7414 | hCoV-19/England/ALDP-B5E28A/2020\|EPI_ISL_723646\|20 | ···················· | ···················· | ···················· | ···················· | ···················· | ···················· |
| X | 7415 | hCoV-19/England/ALDP-B5E2C6/2020\|EPI_ISL_723647\|20 | ···················· | ···················· | ···················· | ···················· | ···················· | ···················· |
| X | 7416 | hCoV-19/England/ALDP-B5E2D5/2020\|EPI_ISL_723648\|20 | ···················· | ···················· | ···················· | ···················· | ···················· | ···················· |
| X | 7417 | hCoV-19/England/ALDP-B5E2E4/2020\|EPI_ISL_723649\|20 | ···················· | ···················· | ···················· | ···················· | ···················· | ···A················ |
| X | 7418 | hCoV-19/England/ALDP-B5E34B/2020\|EPI_ISL_723651\|20 | ···················· | ···················· | ···················· | ···················· | ···················· | ···················· |
| X | 7419 | hCoV-19/England/ALDP-B5E35A/2020\|EPI_ISL_723652\|20 | ···················· | ···················· | ···················· | ···················· | ···················· | ···················· |
| X | 7420 | hCoV-19/England/ALDP-B5E369/2020\|EPI_ISL_723653\|20 | ···················· | ···················· | ···················· | ···················· | ···················· | ···················· |
| X | 7421 | hCoV-19/England/ALDP-B5E387/2020\|EPI_ISL_723654\|20 | ···················· | ···················· | ···················· | ···················· | ···················· | ···················· |
| X | 7422 | hCoV-19/England/ALDP-B5E396/2020\|EPI_ISL_723655\|20 | ···················· | ···················· | ···················· | ···················· | ···················· | ···················· |
| X | 7423 | hCoV-19/England/ALDP-B5E40C/2020\|EPI_ISL_723657\|20 | ···················· | ···················· | ···················· | ···················· | ···················· | ···················· |
| X | 7424 | hCoV-19/England/ALDP-B5E475/2020\|EPI_ISL_723658\|20 | ···················· | ···················· | ···················· | ···················· | ···················· | ···················· |
| X | 7425 | hCoV-19/Denmark/DCGC-14242/2020\|EPI_ISL_711648\|202 | ···················· | ···················· | ···················· | ···················· | ···················· | ···················· |
| X | 7426 | hCoV-19/Denmark/DCGC-14244/2020\|EPI_ISL_711650\|202 | ···················· | ···················· | ···················· | ···················· | ···················· | ···················· |
| X | 7427 | hCoV-19/Denmark/DCGC-14246/2020\|EPI_ISL_711651\|202 | ···················· | ···················· | ···················· | ···················· | ···················· | ···················· |
| X | 7428 | hCoV-19/Denmark/DCGC-14247/2020\|EPI_ISL_711652\|202 | ···················· | ···················· | ···················· | ···················· | ···················· | ···················· |
| X | 7429 | hCoV-19/Denmark/DCGC-14248/2020\|EPI_ISL_711653\|202 | ···················· | ···················· | ···················· | ···················· | ···················· | ···················· |
| X | 7430 | hCoV-19/Denmark/DCGC-14249/2020\|EPI_ISL_711654\|202 | ···················· | ···················· | ···················· | ···················· | ···················· | ···················· |
| X | 7431 | hCoV-19/Denmark/DCGC-14252/2020\|EPI_ISL_711655\|202 | ···················· | ···················· | ···················· | ···················· | ···················· | ···················· |
| X | 7432 | hCoV-19/Denmark/DCGC-14253/2020\|EPI_ISL_711656\|202 | ···················· | ···················· | ···················· | ···················· | ···················· | ···················· |
| X | 7433 | hCoV-19/Denmark/DCGC-14254/2020\|EPI_ISL_711657\|202 | ···················· | ···················· | ···················· | ···················· | ···················· | ···················· |
| X | 7434 | hCoV-19/Denmark/DCGC-14301/2020\|EPI_ISL_711680\|202 | ···················· | ···················· | ···················· | ···················· | ···················· | ···················· |
| X | 7435 | hCoV-19/Denmark/DCGC-14304/2020\|EPI_ISL_711681\|202 | ···················· | ···················· | ···················· | ···················· | ···················· | ···················· |
| X | 7436 | hCoV-19/Denmark/DCGC-14305/2020\|EPI_ISL_711682\|202 | ···················· | ···················· | ···················· | ···················· | ···················· | ···················· |
| X | 7437 | hCoV-19/Denmark/DCGC-14306/2020\|EPI_ISL_711683\|202 | ···················· | ···················· | ···················· | ···················· | ···················· | ···················· |
| X | 7438 | hCoV-19/Denmark/DCGC-14308/2020\|EPI_ISL_711684\|202 | ···················· | ···················· | ···················· | ···················· | ···················· | ···················· |
| X | 7439 | hCoV-19/Denmark/DCGC-14309/2020\|EPI_ISL_711685\|202 | ···················· | ···················· | ···················· | ···················· | ···················· | ···················· |
| X | 7440 | hCoV-19/Denmark/DCGC-14310/2020\|EPI_ISL_711686\|202 | ···················· | ·A·················· | ···················· | ···················· | ···················· | ···A················ |
| X | 7441 | hCoV-19/Denmark/DCGC-14311/2020\|EPI_ISL_711687\|202 | ···················· | ····A··············· | ···················· | ···················· | ···················· | ···················· |
| X | 7442 | hCoV-19/Denmark/DCGC-14312/2020\|EPI_ISL_711688\|202 | ···················· | ···················· | ···················· | ···················· | ···················· | ···················· |
| X | 7443 | hCoV-19/Denmark/DCGC-14313/2020\|EPI_ISL_711689\|202 | ···················· | ···················· | ···················· | ···················· | ···················· | ···················· |
| X | 7444 | hCoV-19/Denmark/DCGC-14314/2020\|EPI_ISL_711690\|202 | ···················· | ···················· | ···················· | ···················· | ···················· | ···················· |
| X | 7445 | hCoV-19/Denmark/DCGC-14317/2020\|EPI_ISL_711691\|202 | ···················· | ···················· | ···················· | ···················· | ···················· | ···················· |
| X | 7446 | hCoV-19/Denmark/DCGC-14319/2020\|EPI_ISL_711692\|202 | ···················· | ···················· | ···················· | ···················· | ···················· | ···················· |
| X | 7447 | hCoV-19/Denmark/DCGC-14322/2020\|EPI_ISL_711693\|202 | ···················· | ···················· | ···················· | ···················· | ···················· | ···················· |
| X | 7448 | hCoV-19/Denmark/DCGC-14323/2020\|EPI_ISL_711694\|202 | ···················· | ···················· | ···················· | ···················· | ···················· | ···················· |
| X | 7449 | hCoV-19/Denmark/DCGC-14324/2020\|EPI_ISL_711695\|202 | ···················· | ···················· | ···················· | ···················· | ···················· | ···················· |
| X | 7450 | hCoV-19/Denmark/DCGC-14326/2020\|EPI_ISL_711696\|202 | ···················· | ···················· | ···················· | ···················· | ···················· | ···················· |
| X | 7451 | hCoV-19/Denmark/DCGC-14327/2020\|EPI_ISL_711697\|202 | ···················· | ···················· | ···················· | ···················· | ···················· | ···················· |
| X | 7452 | hCoV-19/Denmark/DCGC-14328/2020\|EPI_ISL_711698\|202 | ···················· | ···················· | ···················· | ···················· | ···················· | ···················· |
| X | 7453 | hCoV-19/Denmark/DCGC-14329/2020\|EPI_ISL_711699\|202 | ···················· | ···················· | ···················· | ···················· | ···················· | ···················· |
| X | 7454 | hCoV-19/Denmark/DCGC-13718/2020\|EPI_ISL_712980\|202 | ···················· | ···················· | ···················· | ···················· | ···················· | ···················· |
| X | 7455 | hCoV-19/Denmark/DCGC-13719/2020\|EPI_ISL_712981\|202 | ···················· | ···················· | ···················· | ···················· | ···················· | ···················· |
| X | 7456 | hCoV-19/Denmark/DCGC-13720/2020\|EPI_ISL_712982\|202 | ···················· | ···················· | ···················· | ···················· | ···················· | ···················· |
| X | 7457 | hCoV-19/Denmark/DCGC-13721/2020\|EPI_ISL_712983\|202 | ···················· | ···················· | ···················· | ···················· | ···················· | ···················· |
| X | 7458 | hCoV-19/Denmark/DCGC-13723/2020\|EPI_ISL_712984\|202 | ···················· | ···················· | ···················· | ···················· | ···················· | ···················· |
| X | 7459 | hCoV-19/Denmark/DCGC-13725/2020\|EPI_ISL_712985\|202 | ···················· | ···················· | ···················· | ···················· | ···················· | ···················· |
| X | 7460 | hCoV-19/Denmark/DCGC-13728/2020\|EPI_ISL_712986\|202 | ···················· | ···················· | ···················· | ···················· | ···················· | ···················· |
| X | 7461 | hCoV-19/Denmark/DCGC-13730/2020\|EPI_ISL_712987\|202 | ···················· | ···················· | ···················· | ···················· | ···················· | ···················· |
| X | 7462 | hCoV-19/Denmark/DCGC-13733/2020\|EPI_ISL_712988\|202 | ···················· | ···················· | ···················· | ···················· | ···················· | ···················· |
| X | 7463 | hCoV-19/Brazil/RJ-00402/2020\|EPI_ISL_717806\|2020-0 | ···················· | ···················· | ···················· | ···················· | ···················· | ···················· |
| X | 7464 | hCoV-19/Brazil/RJ-00404/2020\|EPI_ISL_717807\|2020-1 | ···················· | ···················· | ···················· | ···················· | ···················· | ···················· |
| X | 7465 | hCoV-19/Brazil/RJ-00405/2020\|EPI_ISL_717808\|2020-1 | ···················· | ···················· | ···················· | ···················· | ···················· | ···················· |
| X | 7466 | hCoV-19/Brazil/RJ-00407/2020\|EPI_ISL_717809\|2020-0 | ···················· | ···················· | ···················· | ···················· | ···················· | ······G············· |
| X | 7467 | hCoV-19/Brazil/RJ-00408/2020\|EPI_ISL_717810\|2020-0 | ···················· | ···················· | ···················· | ···················· | ···················· | ···················· |
| X | 7468 | hCoV-19/Brazil/RJ-00409/2020\|EPI_ISL_717811\|2020-0 | ···················· | ···················· | ···················· | ···················· | ···················· | ···················· |
| X | 7469 | hCoV-19/Brazil/RJ-00410/2020\|EPI_ISL_717812\|2020-0 | ···················· | ···················· | ···················· | ···················· | ···················· | ···················· |
| X | 7470 | hCoV-19/Brazil/RJ-00411/2020\|EPI_ISL_717813\|2020-0 | ···················· | ···················· | ···················· | ···················· | ···················· | ···················· |
| X | 7471 | hCoV-19/Brazil/RJ-00412/2020\|EPI_ISL_717814\|2020-0 | ···················· | ···················· | ···················· | ···················· | ···················· | ···················· |
| X | 7472 | hCoV-19/Brazil/RJ-00413/2020\|EPI_ISL_717815\|2020-0 | ···················· | ···················· | ···················· | ···················· | ···················· | ···················· |
| X | 7473 | hCoV-19/Brazil/RJ-00414/2020\|EPI_ISL_717816\|2020-0 | ···················· | ···················· | ···················· | ···················· | ···················· | ···················· |
| X | 7474 | hCoV-19/Brazil/RJ-00415/2020\|EPI_ISL_717817\|2020-0 | ···················· | ···················· | ···················· | ···················· | ···················· | ···················· |
| X | 7475 | hCoV-19/Brazil/RJ-00416/2020\|EPI_ISL_717818\|2020-1 | ···················· | ···················· | ···················· | ···················· | ···················· | ···················· |

| | | | | | | | |
|---|---|---|---|---|---|---|---|
| X | 7476 hCoV-19/Brazil/RJ-00417/2020\|EPI_ISL_717819\|2020-1 | ···················· | ···················· | ····················· | ···················· | ···················· | ···················· |
| X | 7477 hCoV-19/Brazil/RJ-00418/2020\|EPI_ISL_717820\|2020-1 | ···················· | ···················· | ····················· | ···················· | ···················· | ···················· |
| X | 7478 hCoV-19/Brazil/RJ-00419/2020\|EPI_ISL_717821\|2020-1 | ···················· | ····A··············· | ····················· | ···················· | ···················· | ···················· |
| X | 7479 hCoV-19/Brazil/RJ-00420/2020\|EPI_ISL_717822\|2020-1 | ···················· | ···················· | ····················· | ···················· | ···················· | ···················· |
| X | 7480 hCoV-19/Brazil/RJ-00421/2020\|EPI_ISL_717823\|2020-1 | ···················· | ···················· | ····················· | ···················· | ···················· | ···················· |
| X | 7481 hCoV-19/Brazil/RJ-00422/2020\|EPI_ISL_717824\|2020-1 | ···················· | ···················· | ····················· | ···················· | ···················· | ···················· |
| X | 7482 hCoV-19/Brazil/RJ-00423/2020\|EPI_ISL_717825\|2020-1 | ···················· | ···················· | ····················· | ···················· | ···················· | ···················· |
| X | 7483 hCoV-19/Brazil/RJ-00424/2020\|EPI_ISL_717826\|2020-1 | ···················· | ···················· | ····················· | ···················· | ···················· | ···················· |
| X | 7484 hCoV-19/Brazil/RJ-00425/2020\|EPI_ISL_717827\|2020-1 | ···················· | ···················· | ····················· | ···················· | ···················· | ···················· |
| X | 7485 hCoV-19/Brazil/RJ-00426/2020\|EPI_ISL_717828\|2020-1 | ···················· | ···················· | ····················· | ···················· | ···················· | ···················· |
| X | 7486 hCoV-19/Brazil/RJ-00427/2020\|EPI_ISL_717829\|2020-1 | ···················· | ···················· | ····················· | ···················· | ···················· | ···················· |
| X | 7487 hCoV-19/Brazil/RJ-00428/2020\|EPI_ISL_717830\|2020-1 | ···················· | ···················· | ····················· | ···················· | ···················· | ···················· |
| X | 7488 hCoV-19/Brazil/RJ-00432/2020\|EPI_ISL_717831\|2020-1 | ···················· | ···················· | ····················· | ···················· | ···················· | ······G············· |
| X | 7489 hCoV-19/Brazil/RJ-00437/2020\|EPI_ISL_717832\|2020-0 | ···················· | ···················· | ····················· | ···················· | ···················· | ···················· |
| X | 7490 hCoV-19/Brazil/RJ-00438/2020\|EPI_ISL_717833\|2020-0 | ···················· | ···················· | ····················· | ···················· | ···················· | ···················· |
| X | 7491 hCoV-19/Brazil/RJ-00439/2020\|EPI_ISL_717834\|2020-0 | ···················· | ···················· | ····················· | ···················· | ···················· | ···················· |
| X | 7492 hCoV-19/Brazil/RJ-00440/2020\|EPI_ISL_717835\|2020-0 | ···················· | ···················· | ····················· | ···················· | ···················· | ···················· |
| X | 7493 hCoV-19/Brazil/RJ-00441/2020\|EPI_ISL_717836\|2020-0 | ···················· | ···················· | ····················· | ···················· | ···················· | ···················· |
| X | 7494 hCoV-19/Brazil/RJ-00442/2020\|EPI_ISL_717837\|2020-0 | ···················· | ···················· | ····················· | ···················· | ···················· | ···················· |
| X | 7495 hCoV-19/Brazil/RJ-00443/2020\|EPI_ISL_717838\|2020-0 | ···················· | ···················· | ····················· | ···················· | ···················· | ···················· |
| X | 7496 hCoV-19/Brazil/RJ-00444/2020\|EPI_ISL_717839\|2020-0 | ···················· | ···················· | ····················· | ···················· | ···················· | ···················· |
| X | 7497 hCoV-19/Brazil/RJ-00445/2020\|EPI_ISL_717840\|2020-0 | ···················· | ···················· | ····················· | ···················· | ···················· | ···················· |
| X | 7498 hCoV-19/Brazil/RJ-00446/2020\|EPI_ISL_717841\|2020-0 | ···················· | ···················· | ····················· | ···················· | ···················· | ···················· |
| X | 7499 hCoV-19/Brazil/RJ-00447/2020\|EPI_ISL_717842\|2020-0 | ···················· | ···················· | ····················· | ···················· | ···················· | ···················· |
| X | 7500 hCoV-19/Brazil/RJ-00448/2020\|EPI_ISL_717843\|2020-0 | ···················· | ···················· | ····················· | ···················· | ···················· | ···················· |
| X | 7501 hCoV-19/Brazil/RJ-00449/2020\|EPI_ISL_717844\|2020-0 | ···················· | ···················· | ····················· | ···················· | ···················· | ···················· |
| X | 7502 hCoV-19/Brazil/RJ-00450/2020\|EPI_ISL_717845\|2020-0 | ···················· | ···················· | ····················· | ···················· | ···················· | ···················· |
| X | 7503 hCoV-19/Brazil/RJ-00451/2020\|EPI_ISL_717846\|2020-0 | ···················· | ···················· | ····················· | ···················· | ···················· | ···················· |
| X | 7504 hCoV-19/Brazil/RJ-00452/2020\|EPI_ISL_717847\|2020-0 | ···················· | ···················· | ····················· | ···················· | ···················· | ···················· |
| X | 7505 hCoV-19/Brazil/RJ-00453/2020\|EPI_ISL_717848\|2020-0 | ···················· | ···················· | ····················· | ···················· | ···················· | ···················· |
| X | 7506 hCoV-19/Brazil/RJ-00454/2020\|EPI_ISL_717849\|2020-0 | ···················· | ···················· | ····················· | ···················· | ···················· | ···················· |
| X | 7507 hCoV-19/Brazil/RJ-00455/2020\|EPI_ISL_717850\|2020-0 | ···················· | ···················· | ····················· | ···················· | ···················· | ···················· |
| X | 7508 hCoV-19/Brazil/RJ-00456/2020\|EPI_ISL_717851\|2020-0 | ···················· | ···················· | ····················· | ···················· | ···················· | ···················· |
| X | 7509 hCoV-19/Brazil/RJ-00457/2020\|EPI_ISL_717852\|2020-0 | ···················· | ···················· | ····················· | ···················· | ···················· | ···················· |
| X | 7510 hCoV-19/Brazil/RJ-00458/2020\|EPI_ISL_717853\|2020-0 | ···················· | ···················· | ····················· | ···················· | ···················· | ···················· |
| X | 7511 hCoV-19/Brazil/RJ-00459/2020\|EPI_ISL_717854\|2020-0 | ···················· | ···················· | ····················· | ···················· | ···················· | ···················· |
| X | 7512 hCoV-19/Brazil/RJ-00460/2020\|EPI_ISL_717855\|2020-0 | ···················· | ···················· | ····················· | ···················· | ···················· | ···················· |
| X | 7513 hCoV-19/Brazil/RJ-00461/2020\|EPI_ISL_717856\|2020-0 | ···················· | ···················· | ····················· | ···················· | ···················· | ···················· |
| X | 7514 hCoV-19/Brazil/RJ-00462/2020\|EPI_ISL_717857\|2020-0 | ···················· | ···················· | ····················· | ···················· | ···················· | ···················· |
| X | 7515 hCoV-19/Brazil/RJ-00463/2020\|EPI_ISL_717858\|2020-0 | ···················· | ···················· | ····················· | ···················· | ···················· | ···················· |
| X | 7516 hCoV-19/Brazil/RJ-00464/2020\|EPI_ISL_717859\|2020-0 | ···················· | ···················· | ····················· | ···················· | ···················· | ···················· |
| X | 7517 hCoV-19/Brazil/RJ-00465/2020\|EPI_ISL_717860\|2020-0 | ···················· | ···················· | ····················· | ···················· | ···················· | ···················· |
| X | 7518 hCoV-19/Brazil/RJ-00466/2020\|EPI_ISL_717861\|2020-0 | ···················· | ···················· | ····················· | ···················· | ···················· | ···················· |
| X | 7519 hCoV-19/Brazil/RJ-00467/2020\|EPI_ISL_717862\|2020-0 | ···················· | ···················· | ····················· | ···················· | ···················· | ···················· |
| X | 7520 hCoV-19/Brazil/RJ-00468/2020\|EPI_ISL_717863\|2020-0 | ···················· | ···················· | ····················· | ···················· | ···················· | ···················· |
| X | 7521 hCoV-19/Brazil/RJ-00469/2020\|EPI_ISL_717864\|2020-0 | ···················· | ···················· | ····················· | ···················· | ···················· | ···················· |
| X | 7522 hCoV-19/Brazil/RJ-00470/2020\|EPI_ISL_717865\|2020-0 | ···················· | ···················· | ····················· | ···················· | ···················· | ···················· |
| X | 7523 hCoV-19/Brazil/RJ-00471/2020\|EPI_ISL_717866\|2020-0 | ···················· | ···················· | ····················· | ···················· | ···················· | ···················· |
| X | 7524 hCoV-19/Brazil/RJ-00472/2020\|EPI_ISL_717867\|2020-0 | ···················· | ···················· | ····················· | ···················· | ···················· | ···················· |
| X | 7525 hCoV-19/Brazil/RJ-00473/2020\|EPI_ISL_717868\|2020-0 | ···················· | ···················· | ····················· | ···················· | ···················· | ···················· |
| X | 7526 hCoV-19/Brazil/RJ-00486/2020\|EPI_ISL_717880\|2020-1 | ···················· | ···················· | ····················· | ···················· | ···················· | ···················· |
| X | 7527 hCoV-19/Brazil/RJ-00487/2020\|EPI_ISL_717881\|2020-1 | ···················· | ···················· | ····················· | ···················· | ···················· | ···················· |
| X | 7528 hCoV-19/Brazil/RJ-00488/2020\|EPI_ISL_717882\|2020-1 | ···················· | ···················· | ····················· | ···················· | ···················· | ···················· |
| X | 7529 hCoV-19/Brazil/RJ-00489/2020\|EPI_ISL_717883\|2020-1 | ···················· | ···················· | ····················· | ···················· | ···················· | ···················· |
| X | 7530 hCoV-19/Brazil/RJ-00490/2020\|EPI_ISL_717884\|2020-1 | ···················· | ···················· | ····················· | ···················· | ···················· | ···················· |
| X | 7531 hCoV-19/Brazil/RJ-00491/2020\|EPI_ISL_717885\|2020-1 | ···················· | ···················· | ····················· | ···················· | ···················· | ···················· |
| X | 7532 hCoV-19/Brazil/RJ-00492/2020\|EPI_ISL_717886\|2020-1 | ···················· | ···················· | ····················· | ···················· | ···················· | ···················· |
| X | 7533 hCoV-19/Brazil/RJ-00493/2020\|EPI_ISL_717887\|2020-1 | ···················· | ···················· | ····················· | ···················· | ···················· | ···················· |
| X | 7534 hCoV-19/Brazil/RJ-00494/2020\|EPI_ISL_717888\|2020-1 | ···················· | ···················· | ····················· | ···················· | ···················· | ···················· |
| X | 7535 hCoV-19/Brazil/RJ-00495/2020\|EPI_ISL_717889\|2020-1 | ······C············· | ···················· | ····················· | ···················· | ···················· | ···················· |
| X | 7536 hCoV-19/Brazil/RJ-00496/2020\|EPI_ISL_717890\|2020-1 | ···················· | ···················· | ····················· | ···················· | ···················· | ···················· |
| X | 7537 hCoV-19/Brazil/RJ-00497/2020\|EPI_ISL_717891\|2020-1 | ···················· | ···················· | ····················· | ···················· | ···················· | ···················· |
| X | 7538 hCoV-19/Brazil/RJ-00498/2020\|EPI_ISL_717892\|2020-1 | ········T··········· | ···················· | ····················· | ···················· | ···················· | ···················· |
| X | 7539 hCoV-19/Brazil/RJ-00499/2020\|EPI_ISL_717893\|2020-1 | ···················· | ···················· | ····················· | ···················· | ···················· | ···················· |
| X | 7540 hCoV-19/Brazil/RJ-00500/2020\|EPI_ISL_717894\|2020-1 | ···················· | ···················· | ····················· | ···················· | ···················· | ···················· |
| X | 7541 hCoV-19/Brazil/RJ-00501/2020\|EPI_ISL_717895\|2020-1 | ···················· | ···················· | ····················· | ···················· | ···················· | ···················· |
| X | 7542 hCoV-19/Brazil/RJ-00502/2020\|EPI_ISL_717896\|2020-1 | ···················· | ···················· | ····················· | ···················· | ···················· | ···················· |
| X | 7543 hCoV-19/Brazil/RJ-00503/2020\|EPI_ISL_717897\|2020-1 | ···················· | ···················· | ····················· | ···················· | ···················· | ···················· |
| X | 7544 hCoV-19/Brazil/RJ-00504/2020\|EPI_ISL_717898\|2020-1 | ···················· | ···················· | ····················· | ···················· | ···················· | ···················· |
| X | 7545 hCoV-19/Brazil/RJ-00505/2020\|EPI_ISL_717899\|2020-0 | ···················· | ···················· | ····················· | ···················· | ···················· | ···················· |
| X | 7546 hCoV-19/England/QEUH-B9E1BE/2020\|EPI_ISL_721000\|20 | ···················· | ···················· | ····················· | ···················· | ···················· | ···················· |
| X | 7547 hCoV-19/England/QEUH-B9F37B/2020\|EPI_ISL_721001\|20 | ···················· | ···················· | ····················· | ···················· | ···················· | ···················· |
| X | 7548 hCoV-19/England/QEUH-B9DFF1/2020\|EPI_ISL_721002\|20 | ···················· | ···················· | ····················· | ···················· | ···················· | ···················· |
| X | 7549 hCoV-19/England/QEUH-B9F35D/2020\|EPI_ISL_721003\|20 | ···················· | ···················· | ····················· | ···················· | ···················· | ···················· |
| X | 7550 hCoV-19/England/QEUH-B9E50D/2020\|EPI_ISL_721004\|20 | ···················· | ···················· | ····················· | ···················· | ···················· | ···················· |
| X | 7551 hCoV-19/England/QEUH-BA32D1/2020\|EPI_ISL_721005\|20 | ···················· | ···················· | ····················· | ···················· | ···················· | ···················· |
| X | 7552 hCoV-19/England/QEUH-BA4258/2020\|EPI_ISL_721006\|20 | ···················· | ···················· | ····················· | ···················· | ···················· | ···················· |
| X | 7553 hCoV-19/England/QEUH-B9F7F7/2020\|EPI_ISL_721007\|20 | ···················· | ···················· | ····················· | ···················· | ···················· | ···················· |
| X | 7554 hCoV-19/England/QEUH-B9FB46/2020\|EPI_ISL_721008\|20 | ···················· | ···················· | ····················· | ···················· | ···················· | ···················· |
| X | 7555 hCoV-19/England/QEUH-B9F3D5/2020\|EPI_ISL_721010\|20 | ···················· | ···················· | ····················· | ···················· | ···················· | ···················· |

| | | | | | | | | |
|---|---|---|---|---|---|---|---|---|
| X | 7556 | hCoV-19/England/QEUH-B9F52A/2020\|EPI_ISL_721011\|20 | ···················· | ···················· | ····················· | ···················· | ···················· | ···················· |
| X | 7557 | hCoV-19/England/QEUH-B9F733/2020\|EPI_ISL_721012\|20 | ···················· | ···················· | ····················· | ···················· | ···················· | ···················· |
| X | 7558 | hCoV-19/England/QEUH-B9FE2F/2020\|EPI_ISL_721013\|20 | ···················· | ···················· | ····················· | ···················· | ···················· | ···················· |
| X | 7559 | hCoV-19/England/QEUH-B9F93D/2020\|EPI_ISL_721014\|20 | ···················· | ·A·················· | ····················· | ···················· | ···················· | ···················· |
| X | 7560 | hCoV-19/England/QEUH-B9F45A/2020\|EPI_ISL_721015\|20 | ···················· | ···················· | ····················· | ···················· | ···················· | ···················· |
| X | 7561 | hCoV-19/England/QEUH-BA35D8/2020\|EPI_ISL_721016\|20 | ···················· | ···················· | ····················· | ···················· | ···················· | ···················· |
| X | 7562 | hCoV-19/England/QEUH-B9FA85/2020\|EPI_ISL_721017\|20 | ···················· | ···················· | ····················· | ···················· | ···················· | ···················· |
| X | 7563 | hCoV-19/England/QEUH-B9F9C4/2020\|EPI_ISL_721018\|20 | ···················· | ···················· | ····················· | ···················· | ···················· | ···················· |
| X | 7564 | hCoV-19/England/QEUH-B9E039/2020\|EPI_ISL_721019\|20 | ···················· | ···················· | ····················· | ···················· | ···················· | ···················· |
| X | 7565 | hCoV-19/England/QEUH-B9F3C6/2020\|EPI_ISL_721020\|20 | ···················· | ···················· | ····················· | ···················· | ···················· | ···················· |
| X | 7566 | hCoV-19/England/QEUH-B9E3D6/2020\|EPI_ISL_721021\|20 | ···················· | ···················· | ····················· | ···················· | ···················· | ···················· |
| X | 7567 | hCoV-19/England/QEUH-B9F78E/2020\|EPI_ISL_721022\|20 | ···················· | ···················· | ····················· | ···················· | ···················· | ···················· |
| X | 7568 | hCoV-19/England/QEUH-B9FE10/2020\|EPI_ISL_721023\|20 | ···················· | ···················· | ····················· | ···················· | ···················· | ···················· |
| X | 7569 | hCoV-19/England/QEUH-BA3453/2020\|EPI_ISL_721024\|20 | ···················· | ···················· | ····················· | ···················· | ···················· | ···················· |
| X | 7570 | hCoV-19/England/QEUH-B9FAB2/2020\|EPI_ISL_721025\|20 | ···················· | ···················· | ····················· | ···················· | ···················· | ···················· |
| X | 7571 | hCoV-19/England/QEUH-B9FAA3/2020\|EPI_ISL_721026\|20 | ···················· | ···················· | ····················· | ···················· | ···················· | ···················· |
| X | 7572 | hCoV-19/England/QEUH-BA3C88/2020\|EPI_ISL_721027\|20 | ···················· | ···················· | ····················· | ···················· | ···················· | ···················· |
| X | 7573 | hCoV-19/England/QEUH-B9F40F/2020\|EPI_ISL_721028\|20 | ···················· | ···················· | ····················· | ···················· | ···················· | ···················· |
| X | 7574 | hCoV-19/England/QEUH-BA32C2/2020\|EPI_ISL_721029\|20 | ···················· | ···················· | ····················· | ···················· | ···················· | ···················· |
| X | 7575 | hCoV-19/England/QEUH-B9F979/2020\|EPI_ISL_721030\|20 | ···················· | ···················· | ····················· | ···················· | ···················· | ···················· |
| X | 7576 | hCoV-19/England/QEUH-B9F6DC/2020\|EPI_ISL_721031\|20 | ···················· | ···················· | ····················· | ···················· | ······T············· | ···················· |
| X | 7577 | hCoV-19/England/QEUH-B9F7AC/2020\|EPI_ISL_721033\|20 | ···················· | ···················· | ····················· | ···················· | ···················· | ···················· |
| X | 7578 | hCoV-19/England/QEUH-B9E02A/2020\|EPI_ISL_721034\|20 | ···················· | ···················· | ····················· | ···················· | ···················· | ···················· |
| X | 7579 | hCoV-19/England/QEUH-B9FD8C/2020\|EPI_ISL_721035\|20 | ···················· | ···················· | ····················· | ···················· | ······T············· | ···················· |
| X | 7580 | hCoV-19/England/QEUH-B9F548/2020\|EPI_ISL_721036\|20 | ···················· | ···················· | ····················· | ···················· | ···················· | ···················· |
| X | 7581 | hCoV-19/England/QEUH-B9F742/2020\|EPI_ISL_721039\|20 | ···················· | ···················· | ····················· | ···················· | ···················· | ···················· |
| X | 7582 | hCoV-19/England/QEUH-B9F302/2020\|EPI_ISL_721040\|20 | ···················· | ···················· | ····················· | ···················· | ···················· | ···················· |
| X | 7583 | hCoV-19/England/QEUH-BA35E7/2020\|EPI_ISL_721041\|20 | ···················· | ···················· | ····················· | ···················· | ···················· | ···················· |
| X | 7584 | hCoV-19/England/QEUH-B9F5FD/2020\|EPI_ISL_721042\|20 | ···················· | ···················· | ····················· | ···················· | ···················· | ···················· |
| X | 7585 | hCoV-19/England/QEUH-BA4179/2020\|EPI_ISL_721043\|20 | ···················· | ···················· | ····················· | ···················· | ···················· | ···················· |
| X | 7586 | hCoV-19/England/QEUH-BA3532/2020\|EPI_ISL_721045\|20 | ···················· | ···················· | ····················· | ···················· | ···················· | ···················· |
| X | 7587 | hCoV-19/England/QEUH-B9E01B/2020\|EPI_ISL_721046\|20 | ···················· | ·A·················· | ····················· | ···················· | ···················· | ···················· |
| X | 7588 | hCoV-19/England/QEUH-BA3D58/2020\|EPI_ISL_721047\|20 | ···················· | ···················· | ····················· | ···················· | ···················· | ···················· |
| X | 7589 | hCoV-19/England/QEUH-B9F320/2020\|EPI_ISL_721048\|20 | ···················· | ···················· | ····················· | ···················· | ···················· | ···················· |
| X | 7590 | hCoV-19/England/QEUH-B9F3A8/2020\|EPI_ISL_721049\|20 | ···················· | ···················· | ····················· | ···················· | ······T············· | ···················· |
| X | 7591 | hCoV-19/England/QEUH-B9FEF2/2020\|EPI_ISL_721050\|20 | ···················· | ···················· | ····················· | ···················· | ···················· | ···················· |
| X | 7592 | hCoV-19/England/QEUH-B9F2F6/2020\|EPI_ISL_721051\|20 | ···················· | ···················· | ····················· | ···················· | ···················· | ···················· |
| X | 7593 | hCoV-19/England/QEUH-B9F2BA/2020\|EPI_ISL_721052\|20 | ···················· | ···················· | ····················· | ···················· | ···················· | ···················· |
| X | 7594 | hCoV-19/England/QEUH-BA32FF/2020\|EPI_ISL_721053\|20 | ···················· | ···················· | ····················· | ···················· | ···················· | ···················· |
| X | 7595 | hCoV-19/England/QEUH-BA3550/2020\|EPI_ISL_721054\|20 | ···················· | ···················· | ····················· | ···················· | ···················· | ···················· |
| X | 7596 | hCoV-19/England/QEUH-B9FC8F/2020\|EPI_ISL_721055\|20 | ···················· | ···················· | ····················· | ···················· | ···················· | ···················· |
| X | 7597 | hCoV-19/England/QEUH-B9F84F/2020\|EPI_ISL_721056\|20 | ···················· | ···················· | ····················· | ···················· | ···················· | ···················· |
| X | 7598 | hCoV-19/England/QEUH-B9E093/2020\|EPI_ISL_721057\|20 | ···················· | ···················· | ····················· | ···················· | ···················· | ···················· |
| X | 7599 | hCoV-19/England/QEUH-B9FC43/2020\|EPI_ISL_721058\|20 | ···················· | ···················· | ····················· | ···················· | ···················· | ···················· |
| X | 7600 | hCoV-19/England/QEUH-B9FC52/2020\|EPI_ISL_721059\|20 | ···················· | ···················· | ····················· | ···················· | ···················· | ···················· |
| X | 7601 | hCoV-19/England/QEUH-BA3268/2020\|EPI_ISL_721060\|20 | ···················· | ···················· | ····················· | ···················· | ···················· | ···················· |
| X | 7602 | hCoV-19/England/QEUH-B9E172/2020\|EPI_ISL_721062\|20 | ···················· | ···················· | ····················· | ···················· | ···················· | ···················· |
| X | 7603 | hCoV-19/England/QEUH-BA2D59/2020\|EPI_ISL_721063\|20 | ···················· | ···················· | ····················· | ···················· | ···················· | ···················· |
| X | 7604 | hCoV-19/England/QEUH-B9F5EE/2020\|EPI_ISL_721064\|20 | ···················· | ·A·················· | ····················· | ···················· | ···················· | ···················· |
| X | 7605 | hCoV-19/England/QEUH-BA324A/2020\|EPI_ISL_721065\|20 | ···················· | ···················· | ····················· | ···················· | ···················· | ···················· |
| X | 7606 | hCoV-19/England/QEUH-B9F636/2020\|EPI_ISL_721066\|20 | ···················· | ···················· | ····················· | ···················· | ······T············· | ···················· |
| X | 7607 | hCoV-19/England/QEUH-B9FAD0/2020\|EPI_ISL_721067\|20 | ···················· | ···················· | ····················· | ···················· | ···················· | ···················· |
| X | 7608 | hCoV-19/England/QEUH-B9E0DF/2020\|EPI_ISL_721068\|20 | ···················· | ···················· | ····················· | ···················· | ······T············· | ···················· |
| X | 7609 | hCoV-19/England/QEUH-BA2D68/2020\|EPI_ISL_721069\|20 | ···················· | ···················· | ····················· | ···················· | ···················· | ···················· |
| X | 7610 | hCoV-19/England/QEUH-BA3CD3/2020\|EPI_ISL_721070\|20 | ···················· | ···················· | ····················· | ···················· | ···················· | ···················· |
| X | 7611 | hCoV-19/England/QEUH-BA34DB/2020\|EPI_ISL_721071\|20 | ···················· | ···················· | ····················· | ···················· | ···················· | ···················· |
| X | 7612 | hCoV-19/England/QEUH-B9E321/2020\|EPI_ISL_721072\|20 | ···················· | ···················· | ····················· | ···················· | ···················· | ···················· |
| X | 7613 | hCoV-19/England/QEUH-B9FD13/2020\|EPI_ISL_721073\|20 | ···················· | ···················· | ····················· | ···················· | ···················· | ···················· |
| X | 7614 | hCoV-19/England/QEUH-B9E3B8/2020\|EPI_ISL_721074\|20 | ···················· | ···················· | ····················· | ···················· | ···················· | ···················· |
| X | 7615 | hCoV-19/England/QEUH-B9FD5F/2020\|EPI_ISL_721075\|20 | ···················· | ···················· | ····················· | ···················· | ···················· | ···················· |
| X | 7616 | hCoV-19/England/QEUH-B9F6CD/2020\|EPI_ISL_721076\|20 | ···················· | ···················· | ····················· | ···················· | ···················· | ···················· |
| X | 7617 | hCoV-19/England/QEUH-B9E154/2020\|EPI_ISL_721077\|20 | ···················· | ···················· | ····················· | ···················· | ···················· | ···················· |
| X | 7618 | hCoV-19/England/QEUH-BA2D77/2020\|EPI_ISL_721078\|20 | ···················· | ···················· | ····················· | ···················· | ···················· | ···················· |
| X | 7619 | hCoV-19/England/QEUH-BA421C/2020\|EPI_ISL_721079\|20 | ···················· | ···················· | ····················· | ···················· | ···················· | ···················· |
| X | 7620 | hCoV-19/England/QEUH-BA356F/2020\|EPI_ISL_721080\|20 | ···················· | ···················· | ····················· | ···················· | ···················· | ···················· |
| X | 7621 | hCoV-19/England/QEUH-BA358D/2020\|EPI_ISL_721081\|20 | ···················· | ···················· | ····················· | ···················· | ···················· | ···················· |
| X | 7622 | hCoV-19/England/QEUH-B9F9B5/2020\|EPI_ISL_721082\|20 | ···················· | ···················· | ····················· | ···················· | ···················· | ···················· |
| X | 7623 | hCoV-19/England/QEUH-BA323B/2020\|EPI_ISL_721083\|20 | ···················· | ···················· | ····················· | ···················· | ···················· | ···················· |
| X | 7624 | hCoV-19/England/QEUH-B9E048/2020\|EPI_ISL_721085\|20 | ···················· | ···················· | ····················· | ···················· | ···················· | ···················· |
| X | 7625 | hCoV-19/England/QEUH-B9F988/2020\|EPI_ISL_721086\|20 | ···················· | ···················· | ····················· | ···················· | ···················· | ···················· |
| X | 7626 | hCoV-19/England/QEUH-BA365D/2020\|EPI_ISL_721087\|20 | ···················· | ···················· | ····················· | ···················· | ···················· | ···················· |
| X | 7627 | hCoV-19/England/QEUH-B9F7BB/2020\|EPI_ISL_721088\|20 | ···················· | ···················· | ····················· | ···················· | ···················· | ···················· |
| X | 7628 | hCoV-19/England/QEUH-B9F478/2020\|EPI_ISL_721089\|20 | ···················· | ···················· | ····················· | ···················· | ···················· | ···················· |
| X | 7629 | hCoV-19/England/QEUH-B9FDD7/2020\|EPI_ISL_721090\|20 | ···················· | ···················· | ····················· | ···················· | ···················· | ···················· |
| X | 7630 | hCoV-19/England/QEUH-B9FCE9/2020\|EPI_ISL_721091\|20 | ···················· | ···················· | ····················· | ···················· | ···················· | ···················· |
| X | 7631 | hCoV-19/England/QEUH-BA3CE2/2020\|EPI_ISL_721092\|20 | ···················· | ···················· | ····················· | ···················· | ···················· | ···················· |
| X | 7632 | hCoV-19/England/QEUH-BA2DB3/2020\|EPI_ISL_721093\|20 | ···················· | ······A············· | ····················· | ···················· | ···················· | ···················· |
| X | 7633 | hCoV-19/England/QEUH-BA413D/2020\|EPI_ISL_721094\|20 | ···················· | ···················· | ····················· | ···················· | ···················· | ···················· |
| X | 7634 | hCoV-19/England/QEUH-B9E145/2020\|EPI_ISL_721095\|20 | ···················· | ···················· | ····················· | ···················· | ···················· | ···················· |
| X | 7635 | hCoV-19/England/QEUH-B9F4E1/2020\|EPI_ISL_721096\|20 | ···················· | ·········T·········· | ····················· | ···················· | ···················· | ···················· |

| | | | | | | | | | |
|---|---|---|---|---|---|---|---|---|---|
| X | 7636 | hCoV-19/England/QEUH-B9FD04/2020\|EPI_ISL_721097\|20 | ···················· | ···················· | ···················· | ···················· | ···················· | ···················· | |
| X | 7637 | hCoV-19/England/QEUH-B9F469/2020\|EPI_ISL_721098\|20 | ···················· | ···················· | ···················· | ···················· | ···················· | ···················· | |
| X | 7638 | hCoV-19/England/QEUH-BA359C/2020\|EPI_ISL_721099\|20 | ···················· | ···················· | ···················· | ···················· | ···················· | ···············A···· | |
| X | 7639 | hCoV-19/Netherlands/ZH-EMC-1053/2020\|EPI_ISL_72237 | ···················· | ···················· | ···················· | ···················· | ······T············· | ···················· | |
| X | 7640 | hCoV-19/Netherlands/UT-EMC-151/2020\|EPI_ISL_722371 | ···················· | ···················· | ···················· | ···················· | ···················· | ···················· | |
| X | 7641 | hCoV-19/England/ALDP-B5E035/2020\|EPI_ISL_723626\|20 | ···················· | ···················· | ···················· | ···················· | ···················· | ···················· | |
| X | 7642 | hCoV-19/England/ALDP-B5E080/2020\|EPI_ISL_723629\|20 | ···················· | ···················· | ···················· | ···················· | ···················· | ···················· | |
| X | 7643 | hCoV-19/England/ALDP-B5E484/2020\|EPI_ISL_723659\|20 | ···················· | ···················· | ···················· | ···················· | ···················· | ···················· | |
| X | 7644 | hCoV-19/Wales/ALDP-B5E4C0/2020\|EPI_ISL_723660\|2020 | ···················· | ···················· | ···················· | ···················· | ···················· | ···················· | |
| X | 7645 | hCoV-19/Wales/ALDP-B5E4DF/2020\|EPI_ISL_723661\|2020 | ···················· | ···················· | ···················· | ···················· | ···················· | ···················· | |
| X | 7646 | hCoV-19/England/ALDP-B5E4FD/2020\|EPI_ISL_723662\|20 | ···················· | ···················· | ···················· | ···················· | ···················· | ···················· | |
| X | 7647 | hCoV-19/England/ALDP-B5E509/2020\|EPI_ISL_723663\|20 | ···················· | ···················· | ···················· | ···················· | ···················· | ···················· | |
| X | 7648 | hCoV-19/England/ALDP-B5E518/2020\|EPI_ISL_723664\|20 | ···················· | ···················· | ···················· | ···················· | ···················· | ···················· | |
| X | 7649 | hCoV-19/England/ALDP-B5E527/2020\|EPI_ISL_723665\|20 | ···················· | ···················· | ···················· | ···················· | ···················· | ···················· | |
| X | 7650 | hCoV-19/England/ALDP-B5E536/2020\|EPI_ISL_723666\|20 | ···················· | ···················· | ···················· | ···················· | ···················· | ···················· | |
| X | 7651 | hCoV-19/England/ALDP-B5E563/2020\|EPI_ISL_723667\|20 | ···················· | ···················· | ···················· | ···················· | ···················· | ···················· | |
| X | 7652 | hCoV-19/England/ALDP-B5E5BE/2020\|EPI_ISL_723669\|20 | ···················· | ·A·················· | ···················· | ···················· | ···················· | ···A················ | |
| X | 7653 | hCoV-19/England/ALDP-B5E5CD/2020\|EPI_ISL_723670\|20 | ···················· | ···················· | ···················· | ···················· | ···················· | ···················· | |
| X | 7654 | hCoV-19/England/ALDP-B5E5FA/2020\|EPI_ISL_723671\|20 | ···················· | ···················· | ···················· | ···················· | ···················· | ···················· | |
| X | 7655 | hCoV-19/Wales/ALDP-B5E615/2020\|EPI_ISL_723672\|2020 | ···················· | ···················· | ···················· | ···················· | ···················· | ···················· | |
| X | 7656 | hCoV-19/Wales/ALDP-B73BC0/2020\|EPI_ISL_723673\|2020 | ···················· | ···················· | ···················· | ···················· | ···················· | ···················· | |
| X | 7657 | hCoV-19/England/ALDP-B7586D/2020\|EPI_ISL_723674\|20 | ···················· | ·········T·········· | ···················· | ···················· | ···················· | ···················· | |
| X | 7658 | hCoV-19/England/ALDP-B7587C/2020\|EPI_ISL_723675\|20 | ···················· | ···················· | ···················· | ···················· | ···················· | ···················· | |
| X | 7659 | hCoV-19/England/ALDP-B758A9/2020\|EPI_ISL_723677\|20 | ···················· | ···················· | ···················· | ···················· | ···················· | ···················· | |
| X | 7660 | hCoV-19/England/ALDP-B758C7/2020\|EPI_ISL_723678\|20 | ···················· | ···················· | ···················· | ···················· | ···················· | ···················· | |
| X | 7661 | hCoV-19/England/ALDP-B758E5/2020\|EPI_ISL_723679\|20 | ···················· | ···················· | ···················· | ···················· | ···················· | ···················· | |
| X | 7662 | hCoV-19/England/ALDP-B75900/2020\|EPI_ISL_723680\|20 | ···················· | ···················· | ···················· | ···················· | ···················· | ···················· | |
| X | 7663 | hCoV-19/England/ALDP-B7591F/2020\|EPI_ISL_723681\|20 | ···················· | ···················· | ···················· | ···················· | ···················· | ···················· | |
| X | 7664 | hCoV-19/England/ALDP-B7592E/2020\|EPI_ISL_723682\|20 | ···················· | ···················· | ···················· | ···················· | ···················· | ···A················ | |
| X | 7665 | hCoV-19/England/ALDP-B7593D/2020\|EPI_ISL_723683\|20 | ···················· | ···················· | ···················· | ···················· | ···················· | ···A················ | |
| X | 7666 | hCoV-19/England/ALDP-B7594C/2020\|EPI_ISL_723684\|20 | ···················· | ···················· | ···················· | ···················· | ···················· | ···················· | |
| X | 7667 | hCoV-19/England/ALDP-B75C25/2020\|EPI_ISL_723685\|20 | ···················· | ···················· | ···················· | ···················· | ···················· | ···················· | |
| X | 7668 | hCoV-19/England/ALDP-B75C34/2020\|EPI_ISL_723686\|20 | ···················· | ···················· | ···················· | ···················· | ···················· | ···················· | |
| X | 7669 | hCoV-19/England/ALDP-B75C43/2020\|EPI_ISL_723687\|20 | ···················· | ···················· | ···················· | ···················· | ···················· | ···················· | |
| X | 7670 | hCoV-19/England/ALDP-B75C52/2020\|EPI_ISL_723688\|20 | ···················· | ···················· | ···················· | ···················· | ···················· | ···················· | |
| X | 7671 | hCoV-19/England/ALDP-B75C61/2020\|EPI_ISL_723689\|20 | ···················· | ···················· | ···················· | ···················· | ···················· | ···················· | |
| X | 7672 | hCoV-19/England/ALDP-B75C70/2020\|EPI_ISL_723690\|20 | ···················· | ···················· | ···················· | ···················· | ···················· | ···················· | |
| X | 7673 | hCoV-19/England/ALDP-B75C8F/2020\|EPI_ISL_723691\|20 | ···················· | ···················· | ···················· | ···················· | ···················· | ···················· | |
| X | 7674 | hCoV-19/England/ALDP-B75C9E/2020\|EPI_ISL_723692\|20 | ···················· | ···················· | ···················· | ···················· | ···················· | ···················· | |
| X | 7675 | hCoV-19/England/ALDP-B75CAD/2020\|EPI_ISL_723693\|20 | ···················· | ···················· | ···················· | ···················· | ···················· | ···················· | |
| X | 7676 | hCoV-19/England/ALDP-B75CBC/2020\|EPI_ISL_723694\|20 | ···················· | ···················· | ···················· | ···················· | ···················· | ···················· | |
| X | 7677 | hCoV-19/England/ALDP-B75CCB/2020\|EPI_ISL_723695\|20 | ···················· | ···················· | ···················· | ···················· | ···················· | ···················· | |
| X | 7678 | hCoV-19/England/ALDP-B75CF8/2020\|EPI_ISL_723697\|20 | ···················· | ···················· | ···················· | ···················· | ···················· | ···················· | |
| X | 7679 | hCoV-19/England/ALDP-B75D13/2020\|EPI_ISL_723698\|20 | ··················T· | ···················· | ···················· | ···················· | ···················· | ···················· | 1 |
| X | 7680 | hCoV-19/England/ALDP-B75D22/2020\|EPI_ISL_723699\|20 | ···················· | ···················· | ···················· | ···················· | ···················· | ···················· | |
| X | 7681 | hCoV-19/Brazil/RJ-00474/2020\|EPI_ISL_717869\|2020-0 | ···················· | ···················· | ···················· | ···················· | ···················· | ···················· | |
| X | 7682 | hCoV-19/Brazil/RJ-00475/2020\|EPI_ISL_717870\|2020-0 | ···················· | ···················· | ···················· | ···················· | ···················· | ···················· | |
| X | 7683 | hCoV-19/Brazil/RJ-00476/2020\|EPI_ISL_717871\|2020-0 | ···················· | ···················· | ···················· | ···················· | ···················· | ···················· | |
| X | 7684 | hCoV-19/Brazil/RJ-00477/2020\|EPI_ISL_717872\|2020-0 | ···················· | ···················· | ···················· | ···················· | ···················· | ···················· | |
| X | 7685 | hCoV-19/Brazil/RJ-00478/2020\|EPI_ISL_717873\|2020-0 | ···················· | ···················· | ···················· | ···················· | ···················· | ···················· | |
| X | 7686 | hCoV-19/Brazil/RJ-00479/2020\|EPI_ISL_717874\|2020-1 | ···········T········ | ···················· | ···················· | ···················· | ···················· | ···················· | |
| X | 7687 | hCoV-19/Brazil/RJ-00480/2020\|EPI_ISL_717875\|2020-1 | ···················· | ···················· | ···················· | ···················· | ···················· | ···················· | |
| X | 7688 | hCoV-19/Brazil/RJ-00482/2020\|EPI_ISL_717876\|2020-1 | ···················· | ···················· | ···················· | ···················· | ···················· | ···················· | |
| X | 7689 | hCoV-19/Brazil/RJ-00483/2020\|EPI_ISL_717877\|2020-1 | ···················· | ···················· | ···················· | ···················· | ···················· | ···················· | |
| X | 7690 | hCoV-19/Brazil/RJ-00484/2020\|EPI_ISL_717878\|2020-1 | ···················· | ···················· | ···················· | ···················· | ···················· | ···················· | |
| X | 7691 | hCoV-19/Brazil/RJ-00485/2020\|EPI_ISL_717879\|2020-1 | ···················· | ···················· | ···················· | ···················· | ···················· | ···················· | |

Group 7 - Dec 2020-Jan 2021

| A | B | C | D | E | F | G | H | I | J | K | L |
|---|---|---|---|---|---|---|---|---|---|---|---|
| **Database** | **Database ID** | **GISAID ID** | **GCTGCAATCGTGCTACAACT** | **TGAACTGTTGCGACTACGTG** | **TTCGGAAGAGACAGGTACGTT** | **CACACAATCGAAGCGCAGTA** | **GCTGGTGCTGCAGCTTATTA** | **AGGGTCAAGTGCACAGTCTA** | **mismatches?** | **mismatches?** | **primer set?** |
| AA | 1 | hCoV-19/Denmark/DCGC-18717/2020\|EPI_ISL_749480\|202 | .................... | .................... | .................... | .................... | .................... | .................... | | | |
| AA | 2 | hCoV-19/Denmark/DCGC-18723/2020\|EPI_ISL_749481\|202 | .................... | .................... | .................... | .................... | .................... | .................... | | | |
| AA | 3 | hCoV-19/Denmark/DCGC-18738/2020\|EPI_ISL_749482\|202 | .................... | .................... | .................... | .................... | .................... | .................... | | | |
| AA | 4 | hCoV-19/Denmark/DCGC-18743/2020\|EPI_ISL_749483\|202 | .................... | .................... | .................... | .................... | .................... | .................... | | | |
| AA | 5 | hCoV-19/Denmark/DCGC-18061/2020\|EPI_ISL_749493\|202 | .................... | .................... | .................... | .................... | .................... | .................... | | | |
| AA | 6 | hCoV-19/Denmark/DCGC-18294/2020\|EPI_ISL_749494\|202 | .................... | .................... | .................... | .................... | .................... | .................... | | | |
| AA | 7 | hCoV-19/Denmark/DCGC-18498/2020\|EPI_ISL_749495\|202 | .................... | .................... | .................... | .................... | .................... | .................... | | | |
| AA | 8 | hCoV-19/Denmark/DCGC-18577/2020\|EPI_ISL_749496\|202 | .................... | .A.................. | .................... | .................... | .................... | ..A................. | | | |
| AA | 9 | hCoV-19/Denmark/DCGC-17461/2020\|EPI_ISL_750466\|202 | .................... | .................... | .................... | .................... | .................... | .................... | | | |
| AA | 10 | hCoV-19/Denmark/DCGC-17463/2020\|EPI_ISL_750467\|202 | .................... | .................... | .................... | .................... | .................... | .................... | | | |
| AA | 11 | hCoV-19/Denmark/DCGC-17470/2020\|EPI_ISL_750468\|202 | .................... | .................... | .................... | .................... | .................... | .................... | | | |
| AA | 12 | hCoV-19/Denmark/DCGC-17473/2020\|EPI_ISL_750469\|202 | .................... | .................... | .................... | .................... | .................... | .................... | | | |
| AA | 13 | hCoV-19/Denmark/DCGC-17494/2020\|EPI_ISL_750470\|202 | .................... | .................... | .................... | .................... | .................... | .................... | | | |
| AA | 14 | hCoV-19/Denmark/DCGC-17502/2020\|EPI_ISL_750471\|202 | .................... | .................... | .................... | .................... | .................... | .................... | | | |
| AA | 15 | hCoV-19/Denmark/DCGC-17507/2020\|EPI_ISL_750472\|202 | .................... | .................... | .................... | .................... | .................... | .................... | | | |
| AA | 16 | hCoV-19/Denmark/DCGC-17531/2020\|EPI_ISL_750473\|202 | .................... | .................... | .................... | .................... | .................... | .................... | | | |
| AA | 17 | hCoV-19/Denmark/DCGC-17535/2020\|EPI_ISL_750474\|202 | .................... | .................... | .................... | .................... | .................... | .................... | | | |
| AA | 18 | hCoV-19/Denmark/DCGC-17563/2020\|EPI_ISL_750475\|202 | .................... | .................... | .................... | .................... | .................... | .................... | | | |
| AA | 19 | hCoV-19/Denmark/DCGC-17608/2020\|EPI_ISL_750476\|202 | .................... | .................... | .................... | .................... | .................... | .................... | | | |
| AA | 20 | hCoV-19/Denmark/DCGC-17632/2020\|EPI_ISL_750477\|202 | .................... | .................... | .................... | .................... | .................... | .................... | | | |
| AA | 21 | hCoV-19/Denmark/DCGC-17636/2020\|EPI_ISL_750478\|202 | .................... | .................... | .................... | .................... | .................... | .................... | | | |
| AA | 22 | hCoV-19/Denmark/DCGC-17642/2020\|EPI_ISL_750479\|202 | .................... | .................... | .................... | .................... | .................... | .................... | | | |
| AA | 23 | hCoV-19/Denmark/DCGC-17649/2020\|EPI_ISL_750480\|202 | .................... | .................... | .................... | .................... | .................... | .................... | | | |
| AA | 24 | hCoV-19/Denmark/DCGC-17669/2020\|EPI_ISL_750481\|202 | .................... | .................... | .................... | .................... | .................... | .................... | | | |
| AA | 25 | hCoV-19/Denmark/DCGC-17680/2020\|EPI_ISL_750482\|202 | .................... | .................... | .................... | .................... | .................... | .................... | | | |
| AA | 26 | hCoV-19/Denmark/DCGC-17716/2020\|EPI_ISL_750483\|202 | .................... | .................... | .................... | .................... | .................... | .................... | | | |
| AA | 27 | hCoV-19/Denmark/DCGC-17721/2020\|EPI_ISL_750484\|202 | .................... | .................... | .................... | .................... | .................... | .................... | | | |
| AA | 28 | hCoV-19/Denmark/DCGC-17737/2020\|EPI_ISL_750485\|202 | .................... | .................... | .................... | .................... | .................... | .................... | | | |
| AA | 29 | hCoV-19/Denmark/DCGC-17040/2020\|EPI_ISL_750494\|202 | .................... | .A.................. | .................... | .................... | .................... | ..A................. | | | |
| AA | 30 | hCoV-19/Denmark/DCGC-17351/2020\|EPI_ISL_750495\|202 | .................... | .A.................. | .................... | .................... | .................... | ..A................. | | | |
| AA | 31 | hCoV-19/Denmark/DCGC-17566/2020\|EPI_ISL_750496\|202 | .................... | .................... | .................... | .................... | .................... | .................... | | | |
| AA | 32 | hCoV-19/Denmark/DCGC-17687/2020\|EPI_ISL_750497\|202 | .................... | .................... | .................... | .................... | .................... | .................... | | | |
| AA | 33 | hCoV-19/USA/NM-DOH-2020617125/2020\|EPI_ISL_751804\| | .................... | .................... | .................... | .................... | .................... | .................... | | | |
| AA | 34 | hCoV-19/USA/NM-DOH-2020616523/2020\|EPI_ISL_751805\| | .................... | .................... | .................... | .................... | .................... | .................... | | | |
| AA | 35 | hCoV-19/USA/NM-DOH-2020616576/2020\|EPI_ISL_751806\| | .................... | .................... | .................... | .................... | .................... | .................... | | | |
| AA | 36 | hCoV-19/USA/NM-DOH-2020617002/2020\|EPI_ISL_751807\| | .................... | .A.................. | .................... | .................... | .................... | .................... | | | |
| AA | 37 | hCoV-19/USA/NM-DOH-2020617169/2020\|EPI_ISL_751808\| | .................... | .................... | .................... | .................... | .................... | .................... | | | |
| AA | 38 | hCoV-19/USA/NM-DOH-2020616233/2020\|EPI_ISL_751810\| | .................... | .................... | .................... | .................... | .................... | .................... | | | |
| AA | 39 | hCoV-19/USA/NM-DOH-2020616240/2020\|EPI_ISL_751811\| | .................... | .................... | .................... | .................... | .................... | .................... | | | |
| AA | 40 | hCoV-19/USA/NM-DOH-2020616315/2020\|EPI_ISL_751812\| | .................... | .................... | .................... | .................... | .................... | .................... | | | |
| AA | 41 | hCoV-19/USA/NM-DOH-2020616318/2020\|EPI_ISL_751813\| | .................... | .................... | .................... | .................... | .................... | .................... | | | |
| AA | 42 | hCoV-19/USA/NM-DOH-2020616319/2020\|EPI_ISL_751814\| | .................... | .................... | .................... | .................... | .................... | .................... | | | |
| AA | 43 | hCoV-19/USA/NM-DOH-2020616321/2020\|EPI_ISL_751815\| | .................... | .................... | .................... | .................... | .................... | .................... | | | |
| AA | 44 | hCoV-19/USA/NM-DOH-2020616323/2020\|EPI_ISL_751816\| | .................... | .................... | .................... | .................... | .................... | .................... | | | |
| AA | 45 | hCoV-19/USA/NM-DOH-2020616343/2020\|EPI_ISL_751817\| | .................... | .A.................. | .................... | .................... | .................... | .................... | | | |
| AA | 46 | hCoV-19/USA/NM-DOH-2020616502/2020\|EPI_ISL_751818\| | .................... | .A.................. | .................... | .................... | .................... | .................... | | | |
| AA | 47 | hCoV-19/USA/NM-DOH-2020616527/2020\|EPI_ISL_751819\| | .................... | .................... | .................... | .................... | .................... | .................... | | | |
| AA | 48 | hCoV-19/USA/NM-DOH-2020616538/2020\|EPI_ISL_751821\| | .................... | .................... | .................... | .................... | .................... | .................... | | | |
| AA | 49 | hCoV-19/USA/NM-DOH-2020616547/2020\|EPI_ISL_751822\| | .................... | .A.................. | .................... | .................... | .................... | .................... | | | |
| AA | 50 | hCoV-19/USA/NM-DOH-2020616659/2020\|EPI_ISL_751823\| | .................... | .................... | .................... | .................... | .................... | .................... | | | |
| AA | 51 | hCoV-19/USA/NM-DOH-2020616663/2020\|EPI_ISL_751824\| | .................... | .................... | .................... | .................... | .................... | .................... | | | |
| AA | 53 | hCoV-19/USA/NM-DOH-2020616679/2020\|EPI_ISL_751826\| | .................... | .................... | .................... | .................... | .................... | .................... | | | |
| AA | 54 | hCoV-19/Germany/DE-BE-ChVir-W1710-6707/2020\|EPI_IS | .................... | .................... | .................... | .................... | .................... | .................... | | | |
| AA | 55 | hCoV-19/Germany/DE-BE-ChVir-W1711-8245/2020\|EPI_IS | .................... | .................... | .................... | .................... | .................... | .................... | | | |
| AA | 56 | hCoV-19/Germany/DE-BE-ChVir-W1712-4302/2020\|EPI_IS | .................... | .................... | .................... | .................... | .................... | .................... | | | |
| AA | 57 | hCoV-19/Germany/DE-BE-ChVir-W1713-6783/2020\|EPI_IS | .................... | .................... | .................... | .................... | .................... | .................... | | | |
| AA | 58 | hCoV-19/Germany/DE-BE-ChVir-W1714-6785/2020\|EPI_IS | .................... | .................... | .................... | .................... | .................... | .................... | | | |
| AA | 59 | hCoV-19/Germany/DE-BE-ChVir-W1715-6702/2020\|EPI_IS | ...........T........ | .................... | .................... | .................... | .................... | .................... | | | |
| AA | 60 | hCoV-19/Germany/DE-BE-ChVir-W1716-6716/2020\|EPI_IS | .................... | .................... | .................... | .................... | .................... | .................... | | | |
| AA | 61 | hCoV-19/Germany/DE-BE-ChVir-W1717-6729/2020\|EPI_IS | .................... | .A.................. | .................... | .................... | .................... | .................... | | | |
| AA | 62 | hCoV-19/Germany/DE-BE-ChVir-W1719-6702/2020\|EPI_IS | .................... | .................... | .................... | .................... | .................... | .................... | | | |
| AA | 63 | hCoV-19/Germany/DE-BE-ChVir-W1720-6773/2020\|EPI_IS | .................... | .................... | .................... | .................... | .................... | .................... | | | |
| AA | 64 | hCoV-19/Germany/DE-BE-ChVir-W1721-6777/2020\|EPI_IS | .................... | ......A............. | .................... | .................... | .................... | .................... | | | |
| AA | 65 | hCoV-19/Germany/DE-BE-ChVir-W1722-6784/2020\|EPI_IS | .................... | .................... | .................... | .................... | .................... | .................... | | | |
| AA | 66 | hCoV-19/Germany/DE-BE-ChVir-W1725-8201/2020\|EPI_IS | .................... | .................... | .................... | .................... | .................... | .................... | | | |
| AA | 67 | hCoV-19/Germany/DE-BE-ChVir-W1726-8250/2020\|EPI_IS | .................... | .................... | .................... | .................... | .................... | .................... | | | |
| AA | 68 | hCoV-19/Germany/DE-BE-ChVir-W1727-8260/2020\|EPI_IS | .................... | .................... | .................... | .................... | .................... | .................... | | | |
| AA | 69 | hCoV-19/Germany/DE-BE-ChVir-W1729-5875/2020\|EPI_IS | .................... | .................... | .................... | .................... | .................... | .................... | | | |
| AA | 70 | hCoV-19/Germany/DE-BE-ChVir-W1731-2742/2020\|EPI_IS | .................... | .................... | .................... | .................... | .................... | .................... | | | |

| | | | | | | | | |
|---|---|---|---|---|---|---|---|---|
| AA | 71 | hCoV-19/Germany/DE-BE-ChVir-W2117-4075/2020\|EPI_IS | ···················· | ·A·················· | ···················· | ···················· | ···················· | ···················· |
| AA | 72 | hCoV-19/Germany/DE-BE-ChVir-W2122-5977/2020\|EPI_IS | ···················· | ···················· | ···················· | ···················· | ···················· | ···················· |
| AA | 73 | hCoV-19/Germany/DE-BE-ChVir-W2123-4456/2020\|EPI_IS | ···················· | ·A·················· | ···················· | ···················· | ···················· | ···················· |
| AA | 74 | hCoV-19/Germany/DE-BE-ChVir-W2124-5942/2020\|EPI_IS | ···················· | ···················· | ···················· | ···················· | ···················· | ···················· |
| AA | 75 | hCoV-19/Germany/DE-BE-ChVir-W2125-4401/2020\|EPI_IS | ···················· | ···················· | ···················· | ···················· | ···················· | ···················· |
| AA | 76 | hCoV-19/Germany/DE-BW-ChVir-W1424-8276/2020\|EPI_IS | ···················· | ···················· | ···················· | ···················· | ···················· | ···················· |
| AA | 77 | hCoV-19/Germany/DE-BW-ChVir-W1441-8265/2020\|EPI_IS | ···················· | ···················· | ···················· | ···················· | ···················· | ···················· |
| AA | 78 | hCoV-19/Germany/DE-BY-ChVir-W1050-5794/2020\|EPI_IS | ···················· | ···················· | ···················· | ···················· | ···················· | ···················· |
| AA | 79 | hCoV-19/Germany/DE-BY-ChVir-W1057-5795/2020\|EPI_IS | ···················· | ···················· | ···················· | ···················· | ···················· | ···················· |
| AA | 80 | hCoV-19/Germany/DE-BY-ChVir-W1058-5732/2020\|EPI_IS | ···················· | ···················· | ···················· | ···················· | ···················· | ···················· |
| AA | 81 | hCoV-19/Germany/DE-BY-ChVir-W1068-5746/2020\|EPI_IS | ···················· | ···················· | ···················· | ···················· | ···················· | ···················· |
| AA | 82 | hCoV-19/Germany/DE-BY-ChVir-W1121-5754/2020\|EPI_IS | ···················· | ···················· | ···················· | ···················· | ···················· | ···················· |
| AA | 83 | hCoV-19/Germany/DE-BY-ChVir-W1122-5753/2020\|EPI_IS | ···················· | ···················· | ···················· | ···················· | ···················· | ···················· |
| AA | 84 | hCoV-19/Singapore/1469/2020\|EPI_ISL_754084\|2020-12 | ···················· | ···················· | ···················· | ···················· | ···················· | ···················· |
| AA | 85 | hCoV-19/Singapore/1470/2020\|EPI_ISL_754085\|2020-12 | ···················· | ···A················ | ···················· | ···················· | ···················· | ···················· |
| AA | 86 | hCoV-19/Singapore/1471/2020\|EPI_ISL_754086\|2020-12 | ···················· | ···················· | ···················· | ···················· | ···················· | ··A················· |
| AA | 87 | hCoV-19/Singapore/1472/2020\|EPI_ISL_754087\|2020-12 | ···················· | ·A·················· | ···················· | ···················· | ···················· | ···················· |
| AA | 88 | hCoV-19/Singapore/1473/2020\|EPI_ISL_754088\|2020-12 | ···················· | ···················· | ···················· | ···················· | ···················· | ···················· |
| AA | 89 | hCoV-19/Singapore/1474/2020\|EPI_ISL_754089\|2020-12 | ···················· | ·A·················· | ···················· | ···················· | ···················· | ···················· |
| AA | 90 | hCoV-19/Singapore/1475/2020\|EPI_ISL_754090\|2020-12 | ···················· | ···················· | ···················· | ···················· | ···················· | ···················· |
| AA | 91 | hCoV-19/Singapore/1476/2020\|EPI_ISL_754091\|2020-12 | ···················· | ···················· | ···················· | ···················· | ···················· | ···················· |
| AA | 92 | hCoV-19/Singapore/1477/2020\|EPI_ISL_754092\|2020-12 | ···················· | ···················· | ···················· | ···················· | ···················· | ···················· |
| AA | 93 | hCoV-19/Singapore/1478/2020\|EPI_ISL_754093\|2020-12 | ···················· | ···················· | ···················· | ···················· | ···················· | ···················· |
| AA | 94 | hCoV-19/Singapore/1479/2020\|EPI_ISL_754094\|2020-12 | ···················· | ···················· | ···················· | ···················· | ···················· | ··A················· |
| AA | 95 | hCoV-19/Singapore/1480/2020\|EPI_ISL_754095\|2020-12 | ···················· | ···················· | ···················· | ···················· | ···················· | ···················· |
| AA | 96 | hCoV-19/Singapore/1481/2020\|EPI_ISL_754096\|2020-12 | ···················· | ·A·················· | ···················· | ···················· | ···················· | ···················· |
| AA | 98 | hCoV-19/Singapore/1483/2020\|EPI_ISL_754098\|2020-12 | ···················· | ···················· | ···················· | ···················· | ···················· | ···················· |
| AA | 99 | hCoV-19/Singapore/1490/2020\|EPI_ISL_754099\|2020-12 | ···················· | ···················· | ···················· | ···················· | ···················· | ···················· |
| AA | 100 | hCoV-19/USA/CA-SEARCH-5573/2020\|EPI_ISL_755300\|202 | ···················· | ·A·················· | ···················· | ···················· | ···················· | ···················· |
| AA | 101 | hCoV-19/Chile/LI-4/2020\|EPI_ISL_755302\|2020-08-20 | ···················· | ···················· | ···················· | ···················· | ···················· | ···················· |
| AA | 102 | hCoV-19/Chile/LI-1-4/2020\|EPI_ISL_755303\|2020-08-0 | ···················· | ···················· | ···················· | ···················· | ···················· | ···················· |
| AA | 103 | hCoV-19/Chile/RM-1405-124/2020\|EPI_ISL_755304\|2020 | ···················· | ···················· | ···················· | ···················· | ···················· | ···················· |
| AA | 104 | hCoV-19/USA/ME-HETL-J0102/2020\|EPI_ISL_755306\|2020 | ···················· | ···················· | ···················· | ···················· | ···················· | ···················· |
| AA | 105 | hCoV-19/USA/ME-HETL-J0103/2020\|EPI_ISL_755307\|2020 | ···················· | ···················· | ···················· | ···················· | ···················· | ···················· |
| AA | 106 | hCoV-19/USA/ME-HETL-J0104/2020\|EPI_ISL_755308\|2020 | ···················· | ···················· | ···················· | ···················· | ···················· | ···················· |
| AA | 107 | hCoV-19/USA/ME-HETL-J0105/2020\|EPI_ISL_755309\|2020 | ···················· | ···················· | ···················· | ···················· | ···················· | ···················· |
| AA | 108 | hCoV-19/USA/ME-HETL-J0107/2020\|EPI_ISL_755311\|2020 | ···················· | ···················· | ···················· | ···················· | ···················· | ···················· |
| AA | 109 | hCoV-19/USA/ME-HETL-J0108/2020\|EPI_ISL_755312\|2020 | ···················· | ···················· | ···················· | ···················· | ···················· | ···················· |
| AA | 110 | hCoV-19/USA/ME-HETL-J0109/2020\|EPI_ISL_755313\|2020 | ···················· | ···················· | ···················· | ···················· | ···················· | ···················· |
| AA | 111 | hCoV-19/USA/ME-HETL-J0110/2020\|EPI_ISL_755314\|2020 | ···················· | ···················· | ···················· | ···················· | ···················· | ···················· |
| AA | 113 | hCoV-19/USA/ME-HETL-J0114/2020\|EPI_ISL_755316\|2020 | ···················· | ···················· | ···················· | ···················· | ···················· | ···················· |
| AA | 114 | hCoV-19/USA/ME-HETL-J0115/2020\|EPI_ISL_755317\|2020 | ···················· | ···················· | ···················· | ···················· | ···················· | ···················· |
| AA | 115 | hCoV-19/USA/ME-HETL-J0117/2020\|EPI_ISL_755319\|2020 | ···················· | ···················· | ···················· | ···················· | ···················· | ···················· |
| AA | 116 | hCoV-19/USA/ME-HETL-J0119/2020\|EPI_ISL_755321\|2020 | ···················· | ···················· | ···················· | ···················· | ···················· | ···················· |
| AA | 117 | hCoV-19/USA/ME-HETL-J0124/2020\|EPI_ISL_755323\|2020 | ···················· | ···················· | ···················· | ···················· | ···················· | ···················· |
| AA | 118 | hCoV-19/USA/ME-HETL-J0125/2020\|EPI_ISL_755324\|2020 | ···················· | ···················· | ···················· | ···················· | ···················· | ···················· |
| AA | 119 | hCoV-19/USA/ME-HETL-J0126/2020\|EPI_ISL_755325\|2020 | ···················· | ···················· | ···················· | ···················· | ···················· | ···················· |
| AA | 120 | hCoV-19/USA/ME-HETL-J0129/2020\|EPI_ISL_755326\|2020 | ···················· | ···················· | ···················· | ···················· | ···················· | ···················· |
| AA | 121 | hCoV-19/USA/ME-HETL-J0130/2020\|EPI_ISL_755327\|2020 | ···················· | ···················· | ···················· | ···················· | ···················· | ···················· |
| AA | 122 | hCoV-19/USA/ME-HETL-J0132/2020\|EPI_ISL_755329\|2020 | ···················· | ···················· | ···················· | ···················· | ···················· | ···················· |
| AA | 123 | hCoV-19/USA/ME-HETL-J0135/2020\|EPI_ISL_755330\|2020 | ···················· | ···················· | ···················· | ···················· | ···················· | ···················· |
| AA | 124 | hCoV-19/USA/ME-HETL-J0136/2020\|EPI_ISL_755331\|2020 | ···················· | ···················· | ···················· | ···················· | ···················· | ···················· |
| AA | 125 | hCoV-19/USA/ME-HETL-J0137/2020\|EPI_ISL_755332\|2020 | ···················· | ···················· | ···················· | ···················· | ···················· | ···················· |
| AA | 126 | hCoV-19/USA/ME-HETL-J0138/2020\|EPI_ISL_755333\|2020 | ···················· | ···················· | ···················· | ···················· | ···················· | ···················· |
| AA | 127 | hCoV-19/USA/ME-HETL-J0140/2020\|EPI_ISL_755334\|2020 | ···················· | ···················· | ···················· | ···················· | ···················· | ···················· |
| AA | 128 | hCoV-19/USA/ME-HETL-J0141/2020\|EPI_ISL_755335\|2020 | ···················· | ···················· | ···················· | ···················· | ···················· | ···················· |
| AA | 129 | hCoV-19/USA/ME-HETL-J0145/2020\|EPI_ISL_755337\|2020 | ···················· | ···················· | ···················· | ···················· | ···················· | ···················· |
| AA | 130 | hCoV-19/USA/ME-HETL-J0146/2020\|EPI_ISL_755338\|2020 | ···················· | ···················· | ···················· | ···················· | ···················· | ···················· |
| AA | 131 | hCoV-19/USA/ME-HETL-J0149/2020\|EPI_ISL_755339\|2020 | ···················· | ···················· | ···················· | ···················· | ···················· | ···················· |
| AA | 132 | hCoV-19/USA/ME-HETL-J0150/2020\|EPI_ISL_755340\|2020 | ···················· | ···················· | ···················· | ···················· | ···················· | ···················· |
| AA | 133 | hCoV-19/USA/ME-HETL-J0155/2020\|EPI_ISL_755341\|2020 | ···················· | ···················· | ···················· | ···················· | ···················· | ···················· |
| AA | 134 | hCoV-19/USA/ME-HETL-J0158/2020\|EPI_ISL_755342\|2020 | ···················· | ·A·················· | ···················· | ···················· | ···················· | ···················· |
| AA | 135 | hCoV-19/USA/ME-HETL-J0159/2020\|EPI_ISL_755343\|2020 | ···················· | ·A·················· | ···················· | ···················· | ···················· | ···················· |
| AA | 136 | hCoV-19/USA/ME-HETL-J0161/2020\|EPI_ISL_755344\|2020 | ···················· | ·A·················· | ···················· | ···················· | ···················· | ···················· |
| AA | 137 | hCoV-19/USA/ME-HETL-J0162/2020\|EPI_ISL_755345\|2020 | ···················· | ···················· | ···················· | ···················· | ···················· | ···················· |
| AA | 138 | hCoV-19/USA/ME-HETL-J0163/2020\|EPI_ISL_755346\|2020 | ···················· | ···················· | ···················· | ···················· | ···················· | ···················· |
| AA | 139 | hCoV-19/USA/ME-HETL-J0165/2020\|EPI_ISL_755347\|2020 | ···················· | ···················· | ···················· | ···················· | ···················· | ···················· |
| AA | 140 | hCoV-19/USA/ME-HETL-J0166/2020\|EPI_ISL_755348\|2020 | ···················· | ···················· | ···················· | ···················· | ···················· | ···················· |
| AA | 141 | hCoV-19/USA/ME-HETL-J0167/2020\|EPI_ISL_755349\|2020 | ···················· | ···················· | ···················· | ···················· | ···················· | ···················· |
| AA | 142 | hCoV-19/USA/ME-HETL-J0170/2020\|EPI_ISL_755352\|2020 | ···················· | ···················· | ···················· | ···················· | ···················· | ···················· |
| AA | 143 | hCoV-19/USA/ME-HETL-J0172/2020\|EPI_ISL_755353\|2020 | ···················· | ···················· | ···················· | ···················· | ···················· | ···················· |
| AA | 144 | hCoV-19/USA/ME-HETL-J0174/2020\|EPI_ISL_755355\|2020 | ···················· | ···················· | ···················· | ···················· | ···················· | ···················· |
| AA | 145 | hCoV-19/USA/ME-HETL-J0175/2020\|EPI_ISL_755356\|2020 | ···················· | ·A·················· | ···················· | ···················· | ···················· | ···················· |
| AA | 146 | hCoV-19/USA/ME-HETL-J0176/2020\|EPI_ISL_755357\|2020 | ···················· | ···················· | ···················· | ···················· | ···················· | ···················· |
| AA | 147 | hCoV-19/USA/ME-HETL-J0178/2020\|EPI_ISL_755358\|2020 | ···················· | ···················· | ···················· | ···················· | ···················· | ···················· |
| AA | 148 | hCoV-19/USA/ME-HETL-J0179/2020\|EPI_ISL_755359\|2020 | ···················· | ···················· | ···················· | ···················· | ···················· | ···················· |
| AA | 149 | hCoV-19/England/ALDP-C8B863/2020\|EPI_ISL_756602\|20 | ···················· | ···················· | ···················· | ···················· | ···················· | ···················· |
| AA | 150 | hCoV-19/England/ALDP-C8B20B/2020\|EPI_ISL_756609\|20 | ···················· | ···················· | ···················· | ···················· | ···················· | ···················· |
| AA | 151 | hCoV-19/England/ALDP-C8B5A8/2020\|EPI_ISL_756612\|20 | ···················· | ···················· | ···················· | ···················· | ···················· | ···················· |
| AA | 152 | hCoV-19/England/ALDP-C88EA0/2020\|EPI_ISL_756616\|20 | ···················· | ···················· | ···················· | ···················· | ···················· | ···················· |

| | | | | | | | | |
|---|---|---|---|---|---|---|---|---|
| AA | 153 | hCoV-19/England/ALDP-C8B47E/2020\|EPI_ISL_756619\|20 | ···················· | ···················· | ····················· | ···················· | ···················· | ···················· |
| AA | 154 | hCoV-19/England/ALDP-C8AC68/2020\|EPI_ISL_756620\|20 | ···················· | ···················· | ····················· | ···················· | ···················· | ···················· |
| AA | 155 | hCoV-19/England/CAMC-C84017/2020\|EPI_ISL_756621\|20 | ···················· | ···················· | ····················· | ···················· | ···················· | ···················· |
| AA | 156 | hCoV-19/England/CAMC-C86103/2020\|EPI_ISL_756628\|20 | ···················· | ···················· | ····················· | ···················· | ···················· | ···················· |
| AA | 157 | hCoV-19/England/ALDP-C895E6/2020\|EPI_ISL_756632\|20 | ···················· | ···················· | ····················· | ···················· | ···················· | ···················· |
| AA | 158 | hCoV-19/England/ALDP-C8B7EE/2020\|EPI_ISL_756639\|20 | ···················· | ···················· | ····················· | ···················· | ···················· | ···················· |
| AA | 159 | hCoV-19/England/ALDP-C8B6B4/2020\|EPI_ISL_756640\|20 | ···················· | ···················· | ····················· | ···················· | ···················· | ···················· |
| AA | 160 | hCoV-19/England/ALDP-C8B432/2020\|EPI_ISL_756647\|20 | ···················· | ···················· | ····················· | ···················· | ···················· | ···················· |
| AA | 161 | hCoV-19/England/ALDP-C8B520/2020\|EPI_ISL_756648\|20 | ···················· | ···················· | ····················· | ···················· | ···················· | ···················· |
| AA | 162 | hCoV-19/England/ALDP-C8B809/2020\|EPI_ISL_756651\|20 | ···················· | ···················· | ····················· | ···················· | ···················· | ···················· |
| AA | 163 | hCoV-19/England/ALDP-C8B827/2020\|EPI_ISL_756657\|20 | ···················· | ···················· | ····················· | ···················· | ···················· | ···················· |
| AA | 164 | hCoV-19/England/ALDP-C8AC3B/2020\|EPI_ISL_756658\|20 | ···················· | ···················· | ····················· | ···················· | ···················· | ···················· |
| AA | 165 | hCoV-19/England/CAMC-C85FCE/2020\|EPI_ISL_756659\|20 | ···················· | ···················· | ····················· | ···················· | ···················· | ···················· |
| AA | 166 | hCoV-19/England/ALDP-C8B818/2020\|EPI_ISL_756663\|20 | ···················· | ···················· | ····················· | ···················· | ···················· | ···················· |
| AA | 167 | hCoV-19/England/CAMC-C86A08/2020\|EPI_ISL_756674\|20 | ···················· | ·A·················· | ····················· | ···················· | ···················· | ···A················ |
| AA | 168 | hCoV-19/England/CAMC-C84EE0/2020\|EPI_ISL_756675\|20 | ···················· | ···················· | ····················· | ···················· | ···················· | ···················· |
| AA | 169 | hCoV-19/England/CAMC-C854FC/2020\|EPI_ISL_756677\|20 | ···················· | ···················· | ····················· | ···················· | ···················· | ···················· |
| AA | 170 | hCoV-19/England/CAMC-C83F39/2020\|EPI_ISL_756680\|20 | ···················· | ···················· | ····················· | ···················· | ···················· | ···················· |
| AA | 171 | hCoV-19/England/ALDP-C8B4F6/2020\|EPI_ISL_756685\|20 | ···················· | ···················· | ····················· | ···················· | ···················· | ···················· |
| AA | 172 | hCoV-19/England/ALDP-C88D2B/2020\|EPI_ISL_756686\|20 | ···················· | ···················· | ····················· | ···················· | ······T············· | ···················· |
| AA | 173 | hCoV-19/Denmark/DCGC-21433/2020\|EPI_ISL_757900\|202 | ···················· | ···················· | ····················· | ···················· | ···················· | ···················· |
| AA | 174 | hCoV-19/Denmark/DCGC-21435/2020\|EPI_ISL_757901\|202 | ···················· | ···················· | ····················· | ···················· | ···················· | ···················· |
| AA | 175 | hCoV-19/Denmark/DCGC-21465/2020\|EPI_ISL_757902\|202 | ···················· | ···················· | ····················· | ···················· | ···················· | ···················· |
| AA | 176 | hCoV-19/Denmark/DCGC-21504/2020\|EPI_ISL_757903\|202 | ···················· | ···················· | ····················· | ···················· | ···················· | ···················· |
| AA | 177 | hCoV-19/Denmark/DCGC-21508/2020\|EPI_ISL_757904\|202 | ···················· | ···················· | ····················· | ···················· | ···················· | ···················· |
| AA | 178 | hCoV-19/Denmark/DCGC-21527/2020\|EPI_ISL_757905\|202 | ···················· | ···················· | ····················· | ···················· | ···················· | ···················· |
| AA | 179 | hCoV-19/Denmark/DCGC-21559/2020\|EPI_ISL_757906\|202 | ···················· | ···················· | ····················· | ···················· | ···················· | ···················· |
| AA | 180 | hCoV-19/Denmark/DCGC-21568/2020\|EPI_ISL_757907\|202 | ···················· | ···················· | ····················· | ···················· | ···················· | ···················· |
| AA | 181 | hCoV-19/Denmark/DCGC-21585/2020\|EPI_ISL_757908\|202 | ···················· | ···················· | ····················· | ···················· | ···················· | ···················· |
| AA | 182 | hCoV-19/Denmark/DCGC-21591/2020\|EPI_ISL_757909\|202 | ···················· | ···················· | ····················· | ···················· | ···················· | ···················· |
| AA | 183 | hCoV-19/Denmark/DCGC-21599/2020\|EPI_ISL_757910\|202 | ···················· | ···················· | ····················· | ···················· | ···················· | ···················· |
| AA | 184 | hCoV-19/Denmark/DCGC-21616/2020\|EPI_ISL_757911\|202 | ···················· | ···················· | ····················· | ···················· | ···················· | ···················· |
| AA | 185 | hCoV-19/Denmark/DCGC-21621/2020\|EPI_ISL_757912\|202 | ···················· | ···················· | ····················· | ···················· | ···················· | ···················· |
| AA | 186 | hCoV-19/Denmark/DCGC-21666/2020\|EPI_ISL_757913\|202 | ···················· | ···················· | ····················· | ···················· | ···················· | ···················· |
| AA | 187 | hCoV-19/Denmark/DCGC-21713/2020\|EPI_ISL_757914\|202 | ···················· | ···················· | ····················· | ···················· | ···················· | ···················· |
| AA | 188 | hCoV-19/Denmark/DCGC-21715/2020\|EPI_ISL_757915\|202 | ···················· | ···················· | ····················· | ···················· | ···················· | ···················· |
| AA | 189 | hCoV-19/Denmark/DCGC-21721/2020\|EPI_ISL_757916\|202 | ···················· | ···················· | ····················· | ···················· | ···················· | ···················· |
| AA | 190 | hCoV-19/Denmark/DCGC-21723/2020\|EPI_ISL_757917\|202 | ···················· | ···················· | ····················· | ···················· | ···················· | ···················· |
| AA | 191 | hCoV-19/Denmark/DCGC-21760/2020\|EPI_ISL_757918\|202 | ···················· | ···················· | ····················· | ···················· | ···················· | ···················· |
| AA | 192 | hCoV-19/Denmark/DCGC-21770/2020\|EPI_ISL_757919\|202 | ···················· | ···················· | ····················· | ···················· | ···················· | ···················· |
| AA | 193 | hCoV-19/Denmark/DCGC-21788/2020\|EPI_ISL_757920\|202 | ···················· | ···················· | ····················· | ···················· | ···················· | ···················· |
| AA | 194 | hCoV-19/Denmark/DCGC-21793/2020\|EPI_ISL_757921\|202 | ···················· | ···················· | ····················· | ···················· | ···················· | ···················· |
| AA | 195 | hCoV-19/Denmark/DCGC-21836/2020\|EPI_ISL_757922\|202 | ···················· | ···················· | ····················· | ···················· | ···················· | ···················· |
| AA | 196 | hCoV-19/Denmark/DCGC-21892/2020\|EPI_ISL_757923\|202 | ···················· | ···················· | ····················· | ···················· | ···················· | ···················· |
| AA | 197 | hCoV-19/Denmark/DCGC-21897/2020\|EPI_ISL_757924\|202 | ···················· | ···················· | ····················· | ···················· | ···················· | ···················· |
| AA | 198 | hCoV-19/Denmark/DCGC-21937/2020\|EPI_ISL_757925\|202 | ···················· | ···················· | ····················· | ···················· | ···················· | ···················· |
| AA | 199 | hCoV-19/Denmark/DCGC-21965/2020\|EPI_ISL_757926\|202 | ···················· | ···················· | ····················· | ···················· | ···················· | ···················· |
| AA | 200 | hCoV-19/Denmark/DCGC-21977/2020\|EPI_ISL_757927\|202 | ···················· | ···················· | ····················· | ···················· | ···················· | ···················· |
| AA | 201 | hCoV-19/Denmark/DCGC-21978/2020\|EPI_ISL_757928\|202 | ···················· | ···················· | ····················· | ···················· | ···················· | ···················· |
| AA | 202 | hCoV-19/Denmark/DCGC-21994/2020\|EPI_ISL_757929\|202 | ···················· | ···················· | ····················· | ···················· | ···················· | ···················· |
| AA | 203 | hCoV-19/Denmark/DCGC-22012/2020\|EPI_ISL_757930\|202 | ···················· | ···················· | ····················· | ···················· | ···················· | ···················· |
| AA | 204 | hCoV-19/Denmark/DCGC-22034/2020\|EPI_ISL_757931\|202 | ···················· | ···················· | ····················· | ···················· | ···················· | ···················· |
| AA | 205 | hCoV-19/Denmark/DCGC-22068/2020\|EPI_ISL_757932\|202 | ···················· | ···················· | ····················· | ···················· | ···················· | ···················· |
| AA | 206 | hCoV-19/Denmark/DCGC-22094/2020\|EPI_ISL_757933\|202 | ···················· | ···················· | ····················· | ···················· | ···················· | ···················· |
| AA | 207 | hCoV-19/Denmark/DCGC-22110/2020\|EPI_ISL_757934\|202 | ···················· | ···················· | ····················· | ···················· | ···················· | ···················· |
| AA | 208 | hCoV-19/Denmark/DCGC-22169/2020\|EPI_ISL_757935\|202 | ···················· | ···················· | ····················· | ···················· | ···················· | ···················· |
| AA | 209 | hCoV-19/Denmark/DCGC-22173/2020\|EPI_ISL_757936\|202 | ···················· | ···················· | ····················· | ···················· | ···················· | ···················· |
| AA | 210 | hCoV-19/Denmark/DCGC-22203/2020\|EPI_ISL_757937\|202 | ···················· | ···················· | ····················· | ···················· | ···················· | ···················· |
| AA | 211 | hCoV-19/Denmark/DCGC-22217/2020\|EPI_ISL_757938\|202 | ···················· | ···················· | ····················· | ···················· | ···················· | ···················· |
| AA | 212 | hCoV-19/Denmark/DCGC-22224/2020\|EPI_ISL_757939\|202 | ···················· | ···················· | ····················· | ···················· | ···················· | ···················· |
| AA | 213 | hCoV-19/Denmark/DCGC-22229/2020\|EPI_ISL_757940\|202 | ···················· | ···················· | ····················· | ···················· | ···················· | ···················· |
| AA | 214 | hCoV-19/Denmark/DCGC-22232/2020\|EPI_ISL_757941\|202 | ···················· | ···················· | ····················· | ···················· | ···················· | ···················· |
| AA | 215 | hCoV-19/Denmark/DCGC-22234/2020\|EPI_ISL_757942\|202 | ···················· | ···················· | ···········T········· | ···················· | ···················· | ···················· |
| AA | 216 | hCoV-19/Denmark/DCGC-22305/2020\|EPI_ISL_757943\|202 | ···················· | ···················· | ····················· | ···················· | ···················· | ···················· |
| AA | 217 | hCoV-19/Denmark/DCGC-22327/2020\|EPI_ISL_757944\|202 | ···················· | ···················· | ····················· | ···················· | ···················· | ···················· |
| AA | 218 | hCoV-19/Denmark/DCGC-21425/2020\|EPI_ISL_757960\|202 | ···················· | ···················· | ····················· | ···················· | ···················· | ···················· |
| AA | 219 | hCoV-19/Denmark/DCGC-21615/2020\|EPI_ISL_757961\|202 | ···················· | ···················· | ····················· | ···················· | ···················· | ···················· |
| AA | 220 | hCoV-19/Denmark/DCGC-21628/2020\|EPI_ISL_757962\|202 | ···················· | ···················· | ····················· | ···················· | ···················· | ···················· |
| AA | 221 | hCoV-19/Denmark/DCGC-21665/2020\|EPI_ISL_757963\|202 | ···················· | ···················· | ····················· | ···················· | ···················· | ···················· |
| AA | 222 | hCoV-19/Denmark/DCGC-21844/2020\|EPI_ISL_757964\|202 | ···················· | ···················· | ····················· | ···················· | ···················· | ···················· |
| AA | 223 | hCoV-19/Denmark/DCGC-21948/2020\|EPI_ISL_757965\|202 | ···················· | ···················· | ····················· | ···················· | ···················· | ···················· |
| AA | 224 | hCoV-19/Denmark/DCGC-21970/2020\|EPI_ISL_757966\|202 | ···················· | ···················· | ····················· | ···················· | ···················· | ···················· |
| AA | 225 | hCoV-19/Denmark/DCGC-21986/2020\|EPI_ISL_757967\|202 | ···················· | ···················· | ····················· | ···················· | ···················· | ···················· |
| AA | 226 | hCoV-19/Denmark/DCGC-22027/2020\|EPI_ISL_757968\|202 | ···················· | ···················· | ····················· | ···················· | ···················· | ···················· |
| AA | 227 | hCoV-19/Denmark/DCGC-22058/2020\|EPI_ISL_757969\|202 | ···················· | ···················· | ····················· | ···················· | ···················· | ···················· |
| AA | 228 | hCoV-19/Denmark/DCGC-22219/2020\|EPI_ISL_757970\|202 | ···················· | ···················· | ····················· | ···················· | ···················· | ···················· |
| AA | 229 | hCoV-19/Denmark/DCGC-22235/2020\|EPI_ISL_757971\|202 | ···················· | ···················· | ····················· | ···················· | ···················· | ···················· |
| AA | 230 | hCoV-19/Denmark/DCGC-22265/2020\|EPI_ISL_757972\|202 | ···················· | ···················· | ····················· | ···················· | ···················· | ···················· |
| AA | 231 | hCoV-19/Denmark/DCGC-22321/2020\|EPI_ISL_757973\|202 | ···················· | ···················· | ····················· | ···················· | ···················· | ···················· |
| AA | 232 | hCoV-19/Denmark/DCGC-21375/2020\|EPI_ISL_757975\|202 | ···················· | ···················· | ····················· | ···················· | ···C················ | ···················· |

| | | | | | | | |
|---|---|---|---|---|---|---|---|
| AA | 233 hCoV-19/Denmark/DCGC-21377/2020\|EPI_ISL_757976\|202 | .................... | .................... | .................... | .................... | .................... | .................... |
| AA | 234 hCoV-19/Denmark/DCGC-21378/2020\|EPI_ISL_757977\|202 | .................... | .................... | .................... | .................... | .................... | .................... |
| AA | 235 hCoV-19/Denmark/DCGC-21387/2020\|EPI_ISL_757978\|202 | .................... | .................... | .................... | .................... | .................... | .................... |
| AA | 236 hCoV-19/Denmark/DCGC-21388/2020\|EPI_ISL_757979\|202 | .................... | .................... | .................... | .................... | .................... | .................... |
| AA | 237 hCoV-19/Denmark/DCGC-21399/2020\|EPI_ISL_757980\|202 | .................... | .................... | .................... | .................... | .................... | .................... |
| AA | 238 hCoV-19/Denmark/DCGC-21411/2020\|EPI_ISL_757981\|202 | .................... | .................... | .................... | .................... | .................... | .................... |
| AA | 239 hCoV-19/Denmark/DCGC-21423/2020\|EPI_ISL_757982\|202 | .................... | .................... | .................... | .................... | .................... | .................... |
| AA | 240 hCoV-19/Denmark/DCGC-21427/2020\|EPI_ISL_757983\|202 | .................... | .................... | .................... | .................... | .................... | .................... |
| AA | 241 hCoV-19/Denmark/DCGC-21439/2020\|EPI_ISL_757984\|202 | .................... | .................... | .................... | .................... | .................... | .................... |
| AA | 242 hCoV-19/Denmark/DCGC-21464/2020\|EPI_ISL_757985\|202 | .................... | .................... | .................... | .................... | .................... | .................... |
| AA | 243 hCoV-19/Denmark/DCGC-21467/2020\|EPI_ISL_757986\|202 | .................... | .................... | .................... | .................... | .................... | .................... |
| AA | 244 hCoV-19/Denmark/DCGC-21479/2020\|EPI_ISL_757987\|202 | .................... | .................... | .................... | .................... | .................... | .................... |
| AA | 245 hCoV-19/Denmark/DCGC-21485/2020\|EPI_ISL_757988\|202 | .................... | .................... | .................... | .................... | .................... | .................... |
| AA | 246 hCoV-19/Denmark/DCGC-21491/2020\|EPI_ISL_757989\|202 | .................... | .................... | .................... | .................... | .................... | .................... |
| AA | 247 hCoV-19/Denmark/DCGC-21503/2020\|EPI_ISL_757990\|202 | .................... | .................... | .................... | .................... | .................... | .................... |
| AA | 248 hCoV-19/Denmark/DCGC-21509/2020\|EPI_ISL_757991\|202 | .................... | .................... | .................... | .................... | .................... | .................... |
| AA | 249 hCoV-19/Denmark/DCGC-21519/2020\|EPI_ISL_757992\|202 | .................... | .................... | .................... | .................... | .................... | .................... |
| AA | 250 hCoV-19/Denmark/DCGC-21545/2020\|EPI_ISL_757993\|202 | .................... | .................... | .................... | .................... | .................... | .................... |
| AA | 251 hCoV-19/Denmark/DCGC-21564/2020\|EPI_ISL_757994\|202 | .................... | .................... | .................... | .................... | .................... | .................... |
| AA | 252 hCoV-19/Denmark/DCGC-21569/2020\|EPI_ISL_757995\|202 | .................... | .................... | .................... | .................... | .................... | .................... |
| AA | 253 hCoV-19/Denmark/DCGC-21587/2020\|EPI_ISL_757996\|202 | .................... | .................... | .................... | .................... | .................... | .................... |
| AA | 254 hCoV-19/Denmark/DCGC-21594/2020\|EPI_ISL_757997\|202 | .................... | .................... | .................... | .................... | .................... | .................... |
| AA | 255 hCoV-19/Denmark/DCGC-21596/2020\|EPI_ISL_757998\|202 | .................... | .................... | .................... | .................... | .................... | .................... |
| AA | 256 hCoV-19/Denmark/DCGC-21605/2020\|EPI_ISL_757999\|202 | .................... | .................... | .................... | .................... | .................... | .................... |
| AA | 257 hCoV-19/Denmark/DCGC-19593/2020\|EPI_ISL_748281\|202 | .................... | .................... | .................... | .................... | .................... | .................... |
| AA | 258 hCoV-19/Denmark/DCGC-18760/2020\|EPI_ISL_748282\|202 | .................... | .................... | .................... | .................... | .................... | .................... |
| AA | 259 hCoV-19/Denmark/DCGC-18770/2020\|EPI_ISL_748283\|202 | .................... | .................... | .................... | .................... | .................... | .................... |
| AA | 260 hCoV-19/Denmark/DCGC-18774/2020\|EPI_ISL_748284\|202 | .................... | .................... | .................... | .................... | .................... | .................... |
| AA | 261 hCoV-19/Denmark/DCGC-18782/2020\|EPI_ISL_748285\|202 | .................... | .................... | .................... | .................... | .................... | .................... |
| AA | 262 hCoV-19/Denmark/DCGC-18783/2020\|EPI_ISL_748286\|202 | .................... | .................... | .................... | .................... | .................... | .................... |
| AA | 263 hCoV-19/Denmark/DCGC-18800/2020\|EPI_ISL_748287\|202 | .................... | .................... | .................... | .................... | .................... | .................... |
| AA | 264 hCoV-19/Denmark/DCGC-18820/2020\|EPI_ISL_748288\|202 | .................... | .................... | .................... | .................... | .................... | .................... |
| AA | 265 hCoV-19/Denmark/DCGC-18831/2020\|EPI_ISL_748289\|202 | .................... | .................... | .................... | .................... | .................... | .................... |
| AA | 266 hCoV-19/Denmark/DCGC-18846/2020\|EPI_ISL_748290\|202 | .................... | .................... | .................... | .................... | .................... | .................... |
| AA | 267 hCoV-19/Denmark/DCGC-18956/2020\|EPI_ISL_748291\|202 | .................... | .................... | .................... | .................... | .................... | .................... |
| AA | 268 hCoV-19/Denmark/DCGC-19022/2020\|EPI_ISL_748292\|202 | .................... | .................... | .................... | .................... | .................... | .................... |
| AA | 269 hCoV-19/Denmark/DCGC-19039/2020\|EPI_ISL_748293\|202 | .................... | .................... | .................... | .................... | .................... | .................... |
| AA | 270 hCoV-19/Denmark/DCGC-19095/2020\|EPI_ISL_748294\|202 | .................... | .................... | .................... | .................... | .................... | .................... |
| AA | 271 hCoV-19/Denmark/DCGC-19119/2020\|EPI_ISL_748295\|202 | .................... | .................... | .................... | .................... | .................... | .................... |
| AA | 272 hCoV-19/Denmark/DCGC-19151/2020\|EPI_ISL_748296\|202 | .................... | .................... | .................... | .................... | .................... | .................... |
| AA | 273 hCoV-19/Denmark/DCGC-19158/2020\|EPI_ISL_748297\|202 | .................... | .................... | .................... | .................... | .................... | .................... |
| AA | 274 hCoV-19/Denmark/DCGC-19168/2020\|EPI_ISL_748298\|202 | .................... | .................... | .................... | .................... | .................... | .................... |
| AA | 275 hCoV-19/Denmark/DCGC-19188/2020\|EPI_ISL_748299\|202 | .................... | .................... | .................... | .................... | .................... | .................... |
| AA | 276 hCoV-19/Denmark/DCGC-18640/2020\|EPI_ISL_749522\|202 | .................... | .................... | .................... | .................... | .................... | .................... |
| AA | 277 hCoV-19/Denmark/DCGC-17751/2020\|EPI_ISL_749542\|202 | .................... | .................... | .................... | .................... | .................... | .................... |
| AA | 278 hCoV-19/Denmark/DCGC-17752/2020\|EPI_ISL_749543\|202 | .................... | .................... | .................... | .................... | .................... | .................... |
| AA | 279 hCoV-19/Denmark/DCGC-17753/2020\|EPI_ISL_749544\|202 | .................... | .................... | .................... | .................... | .................... | .................... |
| AA | 280 hCoV-19/Denmark/DCGC-17754/2020\|EPI_ISL_749545\|202 | .................... | .................... | .................... | .................... | .................... | .................... |
| AA | 281 hCoV-19/Denmark/DCGC-17755/2020\|EPI_ISL_749546\|202 | .................... | .................... | .................... | .................... | .................... | .................... |
| AA | 282 hCoV-19/Denmark/DCGC-17756/2020\|EPI_ISL_749547\|202 | .................... | .................... | .................... | .................... | .................... | .................... |
| AA | 283 hCoV-19/Denmark/DCGC-17758/2020\|EPI_ISL_749548\|202 | .................... | .................... | .................... | .................... | .................... | .................... |
| AA | 284 hCoV-19/Denmark/DCGC-17759/2020\|EPI_ISL_749549\|202 | .................... | .................... | .................... | .................... | .................... | .................... |
| AA | 285 hCoV-19/Denmark/DCGC-17760/2020\|EPI_ISL_749550\|202 | .................... | .................... | .................... | .................... | .................... | .................... |
| AA | 286 hCoV-19/Denmark/DCGC-17761/2020\|EPI_ISL_749551\|202 | .................... | .................... | .................... | .................... | .................... | .................... |
| AA | 287 hCoV-19/Denmark/DCGC-17762/2020\|EPI_ISL_749552\|202 | .................... | .................... | .................... | .................... | .................... | .................... |
| AA | 288 hCoV-19/Denmark/DCGC-17763/2020\|EPI_ISL_749553\|202 | .................... | .................... | .................... | .................... | .................... | .................... |
| AA | 289 hCoV-19/Denmark/DCGC-17765/2020\|EPI_ISL_749554\|202 | .................... | .................... | .................... | .................... | .................... | .................... |
| AA | 290 hCoV-19/Denmark/DCGC-17767/2020\|EPI_ISL_749555\|202 | .................... | .................... | .................... | .................... | .................... | .................... |
| AA | 291 hCoV-19/Denmark/DCGC-17768/2020\|EPI_ISL_749556\|202 | .................... | .................... | .................... | .................... | .................... | .................... |
| AA | 292 hCoV-19/Denmark/DCGC-17769/2020\|EPI_ISL_749557\|202 | .................... | .................... | .................... | .................... | .................... | .................... |
| AA | 293 hCoV-19/Denmark/DCGC-17770/2020\|EPI_ISL_749558\|202 | .................... | .................... | .................... | .................... | .................... | .................... |
| AA | 294 hCoV-19/Denmark/DCGC-17771/2020\|EPI_ISL_749559\|202 | .................... | .................... | .................... | .................... | .................... | .................... |
| AA | 295 hCoV-19/Denmark/DCGC-17774/2020\|EPI_ISL_749560\|202 | .................... | .................... | .................... | .................... | .................... | .................... |
| AA | 296 hCoV-19/Denmark/DCGC-17776/2020\|EPI_ISL_749561\|202 | .................... | .................... | .................... | .................... | .................... | .................... |
| AA | 297 hCoV-19/Denmark/DCGC-17777/2020\|EPI_ISL_749562\|202 | .................... | .................... | .................... | .................... | .................... | .................... |
| AA | 298 hCoV-19/Denmark/DCGC-17778/2020\|EPI_ISL_749563\|202 | .................... | .................... | .................... | .................... | .................... | .................... |
| AA | 299 hCoV-19/Denmark/DCGC-17781/2020\|EPI_ISL_749564\|202 | .................... | .................... | .................... | .................... | .................... | .................... |
| AA | 300 hCoV-19/Denmark/DCGC-17782/2020\|EPI_ISL_749565\|202 | .................... | .................... | .................... | .................... | .................... | .................... |
| AA | 301 hCoV-19/Denmark/DCGC-17785/2020\|EPI_ISL_749566\|202 | .................... | .................... | .................... | .................... | .................... | .................... |
| AA | 302 hCoV-19/Denmark/DCGC-17786/2020\|EPI_ISL_749567\|202 | .................... | .................... | .................... | .................... | .................... | .................... |
| AA | 303 hCoV-19/Denmark/DCGC-17788/2020\|EPI_ISL_749568\|202 | .................... | .................... | .................... | .................... | .................... | .................... |
| AA | 304 hCoV-19/Denmark/DCGC-17789/2020\|EPI_ISL_749569\|202 | .................... | .................... | .................... | .................... | .................... | .................... |
| AA | 305 hCoV-19/Denmark/DCGC-17790/2020\|EPI_ISL_749570\|202 | .................... | .................... | .................... | .................... | .................... | .................... |
| AA | 306 hCoV-19/Denmark/DCGC-17791/2020\|EPI_ISL_749571\|202 | .................... | .................... | .................... | .................... | .................... | .................... |
| AA | 307 hCoV-19/Denmark/DCGC-17794/2020\|EPI_ISL_749572\|202 | .................... | .................... | .................... | .................... | .................... | .................... |
| AA | 308 hCoV-19/Denmark/DCGC-17795/2020\|EPI_ISL_749573\|202 | .................... | .................... | .................... | .................... | .................... | .................... |
| AA | 309 hCoV-19/Denmark/DCGC-17797/2020\|EPI_ISL_749574\|202 | .................... | .................... | .................... | .................... | .................... | .................... |
| AA | 310 hCoV-19/Denmark/DCGC-17798/2020\|EPI_ISL_749575\|202 | .................... | .................... | .................... | .................... | .................... | .................... |
| AA | 311 hCoV-19/Denmark/DCGC-17804/2020\|EPI_ISL_749576\|202 | .................... | .................... | .................... | .................... | .................... | .................... |
| AA | 312 hCoV-19/Denmark/DCGC-17805/2020\|EPI_ISL_749577\|202 | .................... | .A.................. | .................... | .................... | .................... | .................... |

| | | | | | | | | | |
|---|---|---|---|---|---|---|---|---|---|
| AA | 313 | hCoV-19/Denmark/DCGC-17806/2020\|EPI_ISL_749578\|202 | .................... | .................... | .................... | .................... | .................... | .................... | |
| AA | 314 | hCoV-19/Denmark/DCGC-17807/2020\|EPI_ISL_749579\|202 | .................... | .................... | .................... | .................... | .................... | .................... | |
| AA | 315 | hCoV-19/Denmark/DCGC-17809/2020\|EPI_ISL_749580\|202 | .................... | .................... | .................... | .................... | .................... | .................... | |
| AA | 316 | hCoV-19/Denmark/DCGC-17811/2020\|EPI_ISL_749581\|202 | .................... | .................... | .................... | .................... | .................... | .................... | |
| AA | 317 | hCoV-19/Denmark/DCGC-17813/2020\|EPI_ISL_749582\|202 | .................... | .................... | .................... | .................... | .................... | .................... | |
| AA | 318 | hCoV-19/Denmark/DCGC-17814/2020\|EPI_ISL_749583\|202 | .................... | .................... | .................... | .................... | .................... | .................... | |
| AA | 319 | hCoV-19/Denmark/DCGC-17815/2020\|EPI_ISL_749584\|202 | .................... | .................... | .................... | .................... | .................... | .................... | |
| AA | 320 | hCoV-19/Denmark/DCGC-17816/2020\|EPI_ISL_749585\|202 | .................... | .................... | .................... | .................... | .................... | .................... | |
| AA | 321 | hCoV-19/Denmark/DCGC-17822/2020\|EPI_ISL_749586\|202 | .................... | .................... | .................... | .................... | .................... | .................... | |
| AA | 322 | hCoV-19/Denmark/DCGC-17826/2020\|EPI_ISL_749587\|202 | .................... | .................... | .................... | .................... | .................... | .................... | |
| AA | 323 | hCoV-19/Denmark/DCGC-17827/2020\|EPI_ISL_749588\|202 | .................... | .................... | .................... | .................... | .................... | .................... | |
| AA | 324 | hCoV-19/Denmark/DCGC-17830/2020\|EPI_ISL_749589\|202 | .................... | .................... | .................... | .................... | .................... | .................... | |
| AA | 325 | hCoV-19/Denmark/DCGC-17831/2020\|EPI_ISL_749590\|202 | .................... | .................... | .................... | .................... | .................... | .................... | |
| AA | 326 | hCoV-19/Denmark/DCGC-17834/2020\|EPI_ISL_749591\|202 | .................... | .................... | .................... | .................... | .................... | .................... | |
| AA | 327 | hCoV-19/Denmark/DCGC-17836/2020\|EPI_ISL_749592\|202 | .................... | .................... | .................... | .................... | .................... | .................... | |
| AA | 328 | hCoV-19/Denmark/DCGC-17838/2020\|EPI_ISL_749593\|202 | .................... | .................... | .................... | .................... | .................... | .................... | |
| AA | 329 | hCoV-19/Denmark/DCGC-17839/2020\|EPI_ISL_749594\|202 | .................... | .................... | .................... | .................... | .................... | .................... | |
| AA | 330 | hCoV-19/Denmark/DCGC-17841/2020\|EPI_ISL_749595\|202 | .................... | .................... | .................... | .................... | .................... | .................... | |
| AA | 331 | hCoV-19/Denmark/DCGC-17842/2020\|EPI_ISL_749596\|202 | .................... | .................... | .................... | .................... | .................... | .................... | |
| AA | 332 | hCoV-19/Denmark/DCGC-17846/2020\|EPI_ISL_749597\|202 | .................... | .................... | .................... | .................... | .................... | .................... | |
| AA | 333 | hCoV-19/Denmark/DCGC-17848/2020\|EPI_ISL_749598\|202 | .................... | .................... | .................... | .................... | .................... | .................... | |
| AA | 334 | hCoV-19/Denmark/DCGC-17851/2020\|EPI_ISL_749599\|202 | .................... | .................... | .................... | .................... | .................... | .................... | |
| AA | 335 | hCoV-19/Denmark/DCGC-17703/2020\|EPI_ISL_750503\|202 | .................... | .................... | .................... | .................... | .................... | .................... | |
| AA | 336 | hCoV-19/Denmark/DCGC-16837/2020\|EPI_ISL_750512\|202 | ..................T. | .................... | .................... | .................... | .................... | .................... | 1 |
| AA | 337 | hCoV-19/Denmark/DCGC-16749/2020\|EPI_ISL_750537\|202 | .................... | .................... | .................... | .................... | .................... | .................... | |
| AA | 338 | hCoV-19/Denmark/DCGC-16750/2020\|EPI_ISL_750538\|202 | .................... | .................... | .................... | .................... | .................... | .................... | |
| AA | 339 | hCoV-19/Denmark/DCGC-16751/2020\|EPI_ISL_750539\|202 | .................... | .................... | .................... | .................... | .................... | .................... | |
| AA | 340 | hCoV-19/Denmark/DCGC-16752/2020\|EPI_ISL_750540\|202 | .................... | .................... | .................... | .................... | .................... | .................... | |
| AA | 341 | hCoV-19/Denmark/DCGC-16753/2020\|EPI_ISL_750541\|202 | .................... | .................... | .................... | .................... | .................... | .................... | |
| AA | 342 | hCoV-19/Denmark/DCGC-16754/2020\|EPI_ISL_750542\|202 | .................... | .................... | .................... | .................... | .................... | .................... | |
| AA | 343 | hCoV-19/Denmark/DCGC-16755/2020\|EPI_ISL_750543\|202 | .................... | .................... | .................... | .................... | .................... | .................... | |
| AA | 344 | hCoV-19/Denmark/DCGC-16756/2020\|EPI_ISL_750544\|202 | .................... | .................... | .................... | .................... | .................... | .................... | |
| AA | 345 | hCoV-19/Denmark/DCGC-16757/2020\|EPI_ISL_750545\|202 | .................... | .................... | .................... | .................... | .................... | .................... | |
| AA | 346 | hCoV-19/Denmark/DCGC-16761/2020\|EPI_ISL_750546\|202 | .................... | .................... | .................... | .................... | .................... | .................... | |
| AA | 347 | hCoV-19/Denmark/DCGC-16762/2020\|EPI_ISL_750547\|202 | .................... | .................... | .................... | .................... | .................... | .................... | |
| AA | 348 | hCoV-19/Denmark/DCGC-16765/2020\|EPI_ISL_750548\|202 | .................... | .................... | .................... | .................... | .................... | .................... | |
| AA | 349 | hCoV-19/Denmark/DCGC-16766/2020\|EPI_ISL_750549\|202 | .................... | .................... | .................... | .................... | .................... | .................... | |
| AA | 350 | hCoV-19/Denmark/DCGC-16767/2020\|EPI_ISL_750550\|202 | .................... | .................... | .................... | .................... | .................... | .................... | |
| AA | 351 | hCoV-19/Denmark/DCGC-16770/2020\|EPI_ISL_750551\|202 | .................... | .................... | .................... | .................... | .................... | .................... | |
| AA | 352 | hCoV-19/Denmark/DCGC-16771/2020\|EPI_ISL_750552\|202 | .................... | .................... | .................... | .................... | .................... | .................... | |
| AA | 353 | hCoV-19/Denmark/DCGC-16772/2020\|EPI_ISL_750553\|202 | .................... | .................... | .................... | .................... | .................... | .................... | |
| AA | 354 | hCoV-19/Denmark/DCGC-16773/2020\|EPI_ISL_750554\|202 | .................... | .................... | .................... | .................... | .................... | .................... | |
| AA | 355 | hCoV-19/Denmark/DCGC-16774/2020\|EPI_ISL_750555\|202 | .................... | .A.................. | .................... | .................... | .................... | .................... | |
| AA | 356 | hCoV-19/Denmark/DCGC-16775/2020\|EPI_ISL_750556\|202 | .................... | .A.................. | .................... | .................... | .................... | ...A................ | |
| AA | 357 | hCoV-19/Denmark/DCGC-16777/2020\|EPI_ISL_750557\|202 | .................... | .................... | .................... | .................... | .................... | .................... | |
| AA | 358 | hCoV-19/Denmark/DCGC-16780/2020\|EPI_ISL_750558\|202 | .................... | .................... | .................... | .................... | .................... | .................... | |
| AA | 359 | hCoV-19/Denmark/DCGC-16782/2020\|EPI_ISL_750559\|202 | .................... | .................... | .................... | .................... | .................... | .................... | |
| AA | 360 | hCoV-19/Denmark/DCGC-16783/2020\|EPI_ISL_750560\|202 | .................... | .................... | .................... | .................... | .................... | .................... | |
| AA | 361 | hCoV-19/Denmark/DCGC-16786/2020\|EPI_ISL_750561\|202 | .................... | .A.................. | .................... | .................... | .................... | .................... | |
| AA | 362 | hCoV-19/Denmark/DCGC-16787/2020\|EPI_ISL_750562\|202 | .................... | .................... | .................... | .................... | .................... | .................... | |
| AA | 363 | hCoV-19/Denmark/DCGC-16788/2020\|EPI_ISL_750563\|202 | .................... | .................... | .................... | .................... | .................... | .................... | |
| AA | 364 | hCoV-19/Denmark/DCGC-16789/2020\|EPI_ISL_750564\|202 | .................... | .................... | .................... | .................... | .................... | .................... | |
| AA | 365 | hCoV-19/Denmark/DCGC-16790/2020\|EPI_ISL_750565\|202 | .................... | .................... | .................... | .................... | .................... | .................... | |
| AA | 366 | hCoV-19/Denmark/DCGC-16792/2020\|EPI_ISL_750566\|202 | .................... | .................... | .................... | .................... | .................... | .................... | |
| AA | 367 | hCoV-19/Denmark/DCGC-16793/2020\|EPI_ISL_750567\|202 | .................... | .................... | .................... | .................... | .................... | .................... | |
| AA | 368 | hCoV-19/Denmark/DCGC-16794/2020\|EPI_ISL_750568\|202 | .................... | .................... | .................... | .................... | .................... | .................... | |
| AA | 369 | hCoV-19/Denmark/DCGC-16795/2020\|EPI_ISL_750569\|202 | .................... | .................... | .................... | .................... | .................... | .................... | |
| AA | 370 | hCoV-19/Denmark/DCGC-16796/2020\|EPI_ISL_750570\|202 | .................... | .................... | .................... | .................... | .................... | .................... | |
| AA | 371 | hCoV-19/Denmark/DCGC-16797/2020\|EPI_ISL_750571\|202 | .................... | .................... | .................... | .................... | .................... | .................... | |
| AA | 372 | hCoV-19/Denmark/DCGC-16798/2020\|EPI_ISL_750572\|202 | .................... | .................... | .................... | .................... | .................... | .................... | |
| AA | 373 | hCoV-19/Denmark/DCGC-16800/2020\|EPI_ISL_750573\|202 | .................... | .................... | .................... | .................... | .................... | .................... | |
| AA | 374 | hCoV-19/Denmark/DCGC-16804/2020\|EPI_ISL_750574\|202 | .................... | .................... | .................... | .................... | .................... | .................... | |
| AA | 375 | hCoV-19/Denmark/DCGC-16807/2020\|EPI_ISL_750575\|202 | .................... | .................... | .................... | .................... | .................... | .................... | |
| AA | 376 | hCoV-19/Denmark/DCGC-16808/2020\|EPI_ISL_750576\|202 | .................... | .................... | .................... | .................... | .................... | .................... | |
| AA | 377 | hCoV-19/Denmark/DCGC-16809/2020\|EPI_ISL_750577\|202 | .................... | .................... | .................... | .................... | .................... | .................... | |
| AA | 378 | hCoV-19/Denmark/DCGC-16811/2020\|EPI_ISL_750578\|202 | .................... | .................... | .................... | .................... | .................... | .................... | |
| AA | 379 | hCoV-19/Denmark/DCGC-16815/2020\|EPI_ISL_750579\|202 | .................... | .................... | .................... | .................... | .................... | .................... | |
| AA | 380 | hCoV-19/Denmark/DCGC-16816/2020\|EPI_ISL_750580\|202 | .................... | .................... | .................... | .................... | .................... | .................... | |
| AA | 381 | hCoV-19/Denmark/DCGC-16817/2020\|EPI_ISL_750581\|202 | .................... | .................... | .................... | .................... | .................... | .................... | |
| AA | 382 | hCoV-19/Denmark/DCGC-16818/2020\|EPI_ISL_750582\|202 | .................... | .................... | .................... | .................... | .................... | .................... | |
| AA | 383 | hCoV-19/Denmark/DCGC-16819/2020\|EPI_ISL_750583\|202 | .................... | .................... | .................... | .................... | .................... | .................... | |
| AA | 384 | hCoV-19/Denmark/DCGC-16821/2020\|EPI_ISL_750584\|202 | .................... | .................... | .................... | .................... | .................... | .................... | |
| AA | 385 | hCoV-19/Denmark/DCGC-16822/2020\|EPI_ISL_750585\|202 | .................... | .................... | .................... | .................... | .................... | .................... | |
| AA | 386 | hCoV-19/Denmark/DCGC-16823/2020\|EPI_ISL_750586\|202 | .................... | .................... | .................... | .................... | .................... | .................... | |
| AA | 387 | hCoV-19/Denmark/DCGC-16824/2020\|EPI_ISL_750587\|202 | .................... | .................... | .................... | .................... | .................... | .................... | |
| AA | 388 | hCoV-19/Denmark/DCGC-16826/2020\|EPI_ISL_750588\|202 | .................... | .................... | .................... | .................... | .................... | .................... | |
| AA | 389 | hCoV-19/Denmark/DCGC-16827/2020\|EPI_ISL_750589\|202 | .................... | .................... | .................... | .................... | .................... | .................... | |
| AA | 390 | hCoV-19/Denmark/DCGC-16829/2020\|EPI_ISL_750590\|202 | .................... | .................... | .................... | .................... | .................... | .................... | |
| AA | 391 | hCoV-19/Denmark/DCGC-16832/2020\|EPI_ISL_750591\|202 | .................... | .................... | .................... | .................... | .................... | .................... | |
| AA | 392 | hCoV-19/Denmark/DCGC-16833/2020\|EPI_ISL_750592\|202 | .................... | .................... | .................... | .................... | .................... | .................... | |

| | | | | | | | |
|---|---|---|---|---|---|---|---|
| AA | 393 hCoV-19/Denmark/DCGC-16834/2020\|EPI_ISL_750593\|202 | .................... | .................... | .................... | .................... | .................... | .................... |
| AA | 394 hCoV-19/Denmark/DCGC-16835/2020\|EPI_ISL_750594\|202 | .................... | .................... | .................... | .................... | .................... | .................... |
| AA | 395 hCoV-19/Denmark/DCGC-16838/2020\|EPI_ISL_750595\|202 | .................... | .................... | .................... | .................... | .................... | .................... |
| AA | 396 hCoV-19/Denmark/DCGC-16841/2020\|EPI_ISL_750596\|202 | .................... | .................... | .................... | .................... | .................... | .................... |
| AA | 397 hCoV-19/Denmark/DCGC-16843/2020\|EPI_ISL_750597\|202 | .................... | .................... | .................... | .................... | .................... | .................... |
| AA | 398 hCoV-19/Denmark/DCGC-16844/2020\|EPI_ISL_750598\|202 | .................... | .................... | .................... | .................... | .................... | .................... |
| AA | 399 hCoV-19/Denmark/DCGC-16847/2020\|EPI_ISL_750599\|202 | .................... | .................... | .................... | .................... | .................... | .................... |
| AA | 400 hCoV-19/USA/NM-DOH-2020616528/2020\|EPI_ISL_751820\| | .................... | .................... | .................... | .................... | .................... | .................... |
| AA | 401 hCoV-19/USA/NM-DOH-2020616689/2020\|EPI_ISL_751827\| | .................... | .................... | .................... | .................... | .................... | .................... |
| AA | 402 hCoV-19/USA/NM-DOH-2020616696/2020\|EPI_ISL_751828\| | .................... | .................... | .................... | .................... | .................... | .................... |
| AA | 403 hCoV-19/USA/NM-DOH-2020616750/2020\|EPI_ISL_751829\| | .................... | .................... | .................... | .................... | .................... | .................... |
| AA | 404 hCoV-19/USA/NM-DOH-2020616766/2020\|EPI_ISL_751830\| | .................... | .................... | .................... | .................... | .................... | .................... |
| AA | 405 hCoV-19/USA/NM-DOH-2020616835/2020\|EPI_ISL_751831\| | .................... | .A.................. | .................... | .................... | .................... | .................... |
| AA | 406 hCoV-19/USA/NM-DOH-2020616848/2020\|EPI_ISL_751832\| | .................... | .A.................. | .................... | .................... | .................... | .................... |
| AA | 407 hCoV-19/USA/NM-DOH-2020616992/2020\|EPI_ISL_751833\| | .................... | .................... | .................... | .................... | .................... | .................... |
| AA | 408 hCoV-19/USA/NM-DOH-2020616994/2020\|EPI_ISL_751834\| | .................... | .................... | .................... | .................... | .................... | .................... |
| AA | 409 hCoV-19/USA/NM-DOH-2020617010/2020\|EPI_ISL_751835\| | .................... | .A.................. | .................... | .................... | .................... | .................... |
| AA | 410 hCoV-19/USA/NM-DOH-2020617018/2020\|EPI_ISL_751836\| | .................... | .................... | .................... | .................... | .................... | .................... |
| AA | 411 hCoV-19/USA/NM-DOH-2020617022/2020\|EPI_ISL_751837\| | .................... | .................... | .................... | .................... | .................... | .................... |
| AA | 412 hCoV-19/USA/NM-DOH-2020617026/2020\|EPI_ISL_751838\| | .................... | .A.................. | .................... | .................... | .................... | .................... |
| AA | 413 hCoV-19/USA/NM-DOH-2020617051/2020\|EPI_ISL_751839\| | .................... | .................... | .................... | .................... | .................... | .................... |
| AA | 414 hCoV-19/USA/NM-DOH-2020617070/2020\|EPI_ISL_751840\| | .................... | .................... | .................... | .................... | .................... | .................... |
| AA | 415 hCoV-19/USA/NM-DOH-2020617082/2020\|EPI_ISL_751841\| | .................... | .A.................. | .................... | .................... | .................... | .................... |
| AA | 416 hCoV-19/USA/NM-DOH-2020617083/2020\|EPI_ISL_751842\| | .................... | .A.................. | .................... | .................... | .................... | .................... |
| AA | 417 hCoV-19/USA/NM-DOH-2020617085/2020\|EPI_ISL_751843\| | .................... | .A.................. | .................... | .................... | .................... | .................... |
| AA | 418 hCoV-19/USA/NM-DOH-2020617119/2020\|EPI_ISL_751844\| | .................... | .A.................. | .................... | .................... | .................... | .................... |
| AA | 419 hCoV-19/USA/NM-DOH-2020617173/2020\|EPI_ISL_751845\| | .................... | .A.................. | .................... | .................... | .................... | .................... |
| AA | 420 hCoV-19/USA/NM-DOH-2020617193/2020\|EPI_ISL_751846\| | .................... | .................... | .................... | .................... | .................... | .................... |
| AA | 421 hCoV-19/Germany/DE-BY-ChVir-W1123-5739/2020\|EPI_IS | .................... | .................... | .................... | .................... | .................... | .................... |
| AA | 422 hCoV-19/Germany/DE-BY-ChVir-W1125-5729/2020\|EPI_IS | .................... | .................... | .................... | .................... | .................... | .................... |
| AA | 423 hCoV-19/Germany/DE-BY-ChVir-W1126-5740/2020\|EPI_IS | .................... | .................... | .................... | .................... | .................... | .................... |
| AA | 424 hCoV-19/Germany/DE-BY-ChVir-W1128-5741/2020\|EPI_IS | .................... | .................... | .................... | .................... | .................... | .................... |
| AA | 425 hCoV-19/Germany/DE-BY-ChVir-W1130-5743/2020\|EPI_IS | .................... | .................... | .................... | .................... | .................... | .................... |
| AA | 426 hCoV-19/Germany/DE-BY-ChVir-W1169-5750/2020\|EPI_IS | .................... | .................... | .................... | .................... | .................... | .................... |
| AA | 427 hCoV-19/Germany/DE-BY-ChVir-W1170-5737/2020\|EPI_IS | .................... | .................... | .................... | .................... | .................... | .................... |
| AA | 428 hCoV-19/Germany/DE-BY-ChVir-W1173-5738/2020\|EPI_IS | .................... | .................... | .................... | .................... | .................... | .................... |
| AA | 429 hCoV-19/Germany/DE-BY-ChVir-W1175-5759/2020\|EPI_IS | .................... | .................... | .................... | .................... | .................... | .................... |
| AA | 430 hCoV-19/Germany/DE-BY-ChVir-W1689-7498/2020\|EPI_IS | .................... | .................... | .................... | .................... | .................... | .................... |
| AA | 431 hCoV-19/Germany/DE-HE-ChVir-W1287-2006/2020\|EPI_IS | .................... | .................... | .................... | .................... | .................... | .................... |
| AA | 432 hCoV-19/Germany/DE-HE-ChVir-W1300-2037/2020\|EPI_IS | .................... | .................... | .................... | .................... | .................... | .................... |
| AA | 433 hCoV-19/Germany/DE-HE-ChVir-W1339-8077/2020\|EPI_IS | .................... | .................... | .................... | .................... | .................... | .................... |
| AA | 434 hCoV-19/Germany/DE-NI-ChVir-W1641-6728/2020\|EPI_IS | .................... | .A.................. | .................... | .................... | .................... | .................... |
| AA | 435 hCoV-19/Germany/DE-NI-ChVir-W1701-6761/2020\|EPI_IS | .................... | .................... | .................... | .................... | .................... | .................... |
| AA | 436 hCoV-19/Germany/DE-NI-ChVir-W1707-6795/2020\|EPI_IS | .................... | .................... | .................... | .................... | .................... | .................... |
| AA | 437 hCoV-19/Germany/DE-NW-ChVir-W1044-5680/2020\|EPI_IS | .................... | .................... | .................... | .................... | .................... | .................... |
| AA | 438 hCoV-19/Germany/DE-NW-ChVir-W1049-5662/2020\|EPI_IS | .................... | .................... | .................... | .................... | .................... | .................... |
| AA | 439 hCoV-19/Germany/DE-SN-ChVir-W1147-4309/2020\|EPI_IS | .................... | .................... | .................... | .................... | .................... | .................... |
| AA | 440 hCoV-19/Germany/DE-SN-ChVir-W1153-4377/2020\|EPI_IS | .................... | .................... | .................... | .................... | .................... | .................... |
| AA | 441 hCoV-19/Germany/DE-SN-ChVir-W1155-4392/2020\|EPI_IS | .................... | .................... | .................... | .................... | .................... | .................... |
| AA | 442 hCoV-19/Germany/DE-SN-ChVir-W1157-4330/2020\|EPI_IS | .................... | .................... | .................... | .................... | .................... | .................... |
| AA | 443 hCoV-19/Germany/DE-SN-ChVir-W1275-4371/2020\|EPI_IS | .................... | .................... | .................... | .................... | .................... | .................... |
| AA | 444 hCoV-19/Germany/DE-SN-ChVir-W1276-4387/2020\|EPI_IS | .................... | .................... | .................... | .................... | .................... | .................... |
| AA | 445 hCoV-19/Germany/DE-ST-ChVir-W1702-6718/2020\|EPI_IS | .................... | .................... | .................... | .................... | .................... | .................... |
| AA | 446 hCoV-19/Germany/DE-BE-ChVir-D697-8137/2020\|EPI_ISL | .................... | .................... | .................... | .................... | .................... | .................... |
| AA | 447 hCoV-19/Germany/DE-BE-ChVir-D750-8181/2020\|EPI_ISL | .................... | .................... | .................... | .................... | .................... | .................... |
| AA | 448 hCoV-19/Germany/DE-BE-ChVir-D751-5650/2020\|EPI_ISL | .................... | .................... | .................... | .................... | .................... | .................... |
| AA | 449 hCoV-19/Germany/DE-BE-ChVir-D754-8111/2020\|EPI_ISL | .................... | .................... | .................... | .................... | .................... | .................... |
| AA | 450 hCoV-19/Bangladesh/SUST-GEB-Sunamganj-4/2020\|EPI_I | .................... | .................... | .................... | .................... | .................... | .................... |
| AA | 451 hCoV-19/Singapore/1498/2020\|EPI_ISL_754107\|2020-12 | .................... | .................... | .................... | .................... | .................... | .................... |
| AA | 452 hCoV-19/Singapore/1499/2020\|EPI_ISL_754108\|2020-12 | .................... | .................... | .................... | .................... | .................... | .................... |
| AA | 453 hCoV-19/Bangladesh/SUST-GEB-Habiganj-1/2020\|EPI_IS | .................... | .................... | .................... | .................... | .................... | .................... |
| AA | 454 hCoV-19/USA/CA-LACPHL-AF00027/2020\|EPI_ISL_754110\| | ..A................. | .................... | .................... | .................... | .................... | .................... |
| AA | 455 hCoV-19/USA/CA-LACPHL-AF00028/2020\|EPI_ISL_754111\| | .................... | .................... | .................... | .................... | .................... | .................... |
| AA | 456 hCoV-19/USA/CA-LACPHL-AF00029/2020\|EPI_ISL_754112\| | .................... | .................... | .................... | .................... | .................... | .................... |
| AA | 457 hCoV-19/USA/CA-LACPHL-AF00030/2020\|EPI_ISL_754113\| | .................... | .................... | .................... | .................... | .................... | .................... |
| AA | 458 hCoV-19/USA/CA-LACPHL-AF00031/2020\|EPI_ISL_754114\| | .................... | .................... | .................... | .................... | .................... | .................... |
| AA | 459 hCoV-19/USA/CA-LACPHL-AF00032/2020\|EPI_ISL_754115\| | .................... | .................... | .................... | .................... | .................... | .................... |
| AA | 460 hCoV-19/USA/CA-LACPHL-AF00033/2020\|EPI_ISL_754116\| | .................... | .................... | .................... | .................... | .................... | .................... |
| AA | 462 hCoV-19/USA/ME-HETL-J0180/2020\|EPI_ISL_755360\|2020 | .................... | .................... | .................... | .................... | .................... | .................... |
| AA | 463 hCoV-19/USA/ME-HETL-J0181/2020\|EPI_ISL_755361\|2020 | .................... | .................... | .................... | .................... | .................... | .................... |
| AA | 464 hCoV-19/USA/ME-HETL-J0184/2020\|EPI_ISL_755362\|2020 | .................... | .................... | .................... | .................... | .................... | .................... |
| AA | 466 hCoV-19/USA/ME-HETL-J0188/2020\|EPI_ISL_755366\|2020 | .................... | .................... | .................... | .................... | .................... | .................... |
| AA | 467 hCoV-19/USA/ME-HETL-J0190/2020\|EPI_ISL_755367\|2020 | .................... | .................... | .................... | .................... | .................... | .................... |
| AA | 470 hCoV-19/USA/ME-HETL-J0199/2020\|EPI_ISL_755370\|2020 | .................... | .................... | .................... | .................... | .................... | .................... |
| AA | 471 hCoV-19/USA/ME-HETL-J0203/2020\|EPI_ISL_755371\|2020 | .................... | .................... | .................... | .................... | .................... | .................... |
| AA | 472 hCoV-19/USA/ME-HETL-J0204/2020\|EPI_ISL_755372\|2020 | .................... | .................... | .................... | .................... | .................... | .................... |
| AA | 473 hCoV-19/USA/ME-HETL-J0206/2020\|EPI_ISL_755373\|2020 | .................... | .................... | .................... | .................... | .................... | .................... |
| AA | 474 hCoV-19/USA/ME-HETL-J0210/2020\|EPI_ISL_755374\|2020 | .................... | .................... | .................... | .................... | .................... | .................... |
| AA | 475 hCoV-19/USA/ME-HETL-J0212/2020\|EPI_ISL_755375\|2020 | .................... | .................... | .................... | .................... | .................... | .................... |
| AA | 476 hCoV-19/USA/ME-HETL-J0213/2020\|EPI_ISL_755376\|2020 | .................... | .................... | .................... | .................... | N................... | .................... |

| | | | | | | | | |
|---|---|---|---|---|---|---|---|---|
| AA | 477 hCoV-19/USA/ME-HETL-J0214/2020\|EPI_ISL_755377\|2020 | ···················· | ···················· | ····················· | ···················· | ···················· | ···················· | |
| AA | 478 hCoV-19/USA/ME-HETL-J0215/2020\|EPI_ISL_755378\|2020 | ···················· | ···················· | ····················· | ···················· | ···················· | ···················· | |
| AA | 479 hCoV-19/USA/ME-HETL-J0216/2020\|EPI_ISL_755379\|2020 | ···················· | ·A·················· | ····················· | ···················· | ···················· | ···················· | |
| AA | 480 hCoV-19/USA/ME-HETL-J0217/2020\|EPI_ISL_755380\|2020 | ··················T· | ···················· | ····················· | ···················· | ···················· | ···················· | 1 |
| AA | 481 hCoV-19/USA/ME-HETL-J0220/2020\|EPI_ISL_755381\|2020 | ···················· | ···················· | ····················· | ···················· | ···················· | ···················· | |
| AA | 482 hCoV-19/USA/ME-HETL-J0221/2020\|EPI_ISL_755382\|2020 | ···················· | ···················· | ····················· | ···················· | ···················· | ···················· | |
| AA | 483 hCoV-19/USA/ME-HETL-J0222/2020\|EPI_ISL_755383\|2020 | ···················· | ···················· | ····················· | ···················· | ···················· | ···················· | |
| AA | 484 hCoV-19/USA/ME-HETL-J0224/2020\|EPI_ISL_755384\|2020 | ···················· | ···················· | ····················· | ···················· | ···················· | ···················· | |
| AA | 485 hCoV-19/USA/ME-HETL-J0226/2020\|EPI_ISL_755386\|2020 | ···················· | ···················· | ····················· | ···················· | ···················· | ···················· | |
| AA | 486 hCoV-19/USA/ME-HETL-J0227/2020\|EPI_ISL_755387\|2020 | ···················· | ·A·················· | ····················· | ···················· | ···················· | ···················· | |
| AA | 487 hCoV-19/USA/ME-HETL-J0228/2020\|EPI_ISL_755388\|2020 | ···················· | ···················· | ····················· | ···················· | ···················· | ···················· | |
| AA | 488 hCoV-19/USA/ME-HETL-J0232/2020\|EPI_ISL_755389\|2020 | ··················T· | ···················· | ····················· | ···················· | ···················· | ···················· | 1 |
| AA | 489 hCoV-19/USA/ME-HETL-J0233/2020\|EPI_ISL_755390\|2020 | ···················· | ···················· | ····················· | ···················· | ···················· | ···················· | |
| AA | 490 hCoV-19/USA/ME-HETL-J0234/2020\|EPI_ISL_755391\|2020 | ···················· | ···················· | ····················· | ···················· | ···················· | ···················· | |
| AA | 491 hCoV-19/USA/ME-HETL-J0235/2020\|EPI_ISL_755392\|2020 | ···················· | ···················· | ····················· | ···················· | ···················· | ···················· | |
| AA | 492 hCoV-19/USA/ME-HETL-J0238/2020\|EPI_ISL_755394\|2020 | ···················· | ···················· | ····················· | ···················· | ···················· | ···················· | |
| AA | 493 hCoV-19/USA/ME-HETL-J0239/2020\|EPI_ISL_755395\|2020 | ···················· | ···················· | ····················· | ···················· | ···················· | ···················· | |
| AA | 494 hCoV-19/USA/ME-HETL-J0240/2020\|EPI_ISL_755396\|2020 | ···················· | ···················· | ····················· | ···················· | ···················· | ···················· | |
| AA | 495 hCoV-19/USA/ME-HETL-J0241/2020\|EPI_ISL_755397\|2020 | ···················· | ···················· | ····················· | ···················· | ···················· | ···················· | |
| AA | 496 hCoV-19/USA/ME-HETL-J0242/2020\|EPI_ISL_755398\|2020 | ···················· | ···················· | ····················· | ···················· | ···················· | ···················· | |
| AA | 497 hCoV-19/USA/ME-HETL-J0243/2020\|EPI_ISL_755399\|2020 | ···················· | ···················· | ····················· | ···················· | ···················· | ···················· | |
| AA | 498 hCoV-19/USA/ME-HETL-J0301/2020\|EPI_ISL_755439\|2020 | ···················· | ···················· | ····················· | ···················· | ···················· | ···················· | |
| AA | 499 hCoV-19/USA/ME-HETL-J0303/2020\|EPI_ISL_755441\|2020 | ···················· | ············C······· | ····················· | ···················· | ···················· | ···················· | |
| AA | 500 hCoV-19/USA/ME-HETL-J0306/2020\|EPI_ISL_755443\|2020 | ···················· | ·A·················· | ····················· | ···················· | ···················· | ···················· | |
| AA | 501 hCoV-19/USA/ME-HETL-J0312/2020\|EPI_ISL_755444\|2020 | ···················· | ···················· | ····················· | ···················· | ···················· | ···················· | |
| AA | 502 hCoV-19/USA/ME-HETL-J0316/2020\|EPI_ISL_755446\|2020 | ···················· | ···················· | ····················· | ···················· | ···················· | ···················· | |
| AA | 503 hCoV-19/USA/ME-HETL-J0318/2020\|EPI_ISL_755448\|2020 | ···················· | ···················· | ····················· | ···················· | ···················· | ···················· | |
| AA | 504 hCoV-19/England/QEUH-C7FEE8/2020\|EPI_ISL_756690\|20 | ···················· | ···················· | ····················· | ···················· | ···················· | ···················· | |
| AA | 505 hCoV-19/England/MILK-C7825C/2020\|EPI_ISL_756692\|20 | ···················· | ···················· | ····················· | ···················· | ······T············· | ···················· | |
| AA | 506 hCoV-19/England/MILK-C78A27/2020\|EPI_ISL_756693\|20 | ···················· | ·A·················· | ····················· | ···················· | ···················· | ···A················ | |
| AA | 507 hCoV-19/England/MILK-C789EE/2020\|EPI_ISL_756694\|20 | ···················· | ···················· | ····················· | ···················· | ···················· | ···················· | |
| AA | 508 hCoV-19/England/MILK-C787D5/2020\|EPI_ISL_756701\|20 | ···················· | ···················· | ····················· | ···················· | ···················· | ···················· | |
| AA | 509 hCoV-19/England/QEUH-C7C249/2020\|EPI_ISL_756702\|20 | ···················· | ···················· | ····················· | ···················· | ···················· | ···················· | |
| AA | 510 hCoV-19/England/MILK-C782C5/2020\|EPI_ISL_756703\|20 | ···················· | ·A·················· | ····················· | ···················· | ···················· | ···A················ | |
| AA | 511 hCoV-19/England/MILK-C7AE65/2020\|EPI_ISL_756704\|20 | ···················· | ···················· | ····················· | ···················· | ······T············· | ···················· | |
| AA | 512 hCoV-19/England/MILK-C7B611/2020\|EPI_ISL_756778\|20 | ···················· | ···················· | ····················· | ···················· | ···················· | ···················· | |
| AA | 513 hCoV-19/Denmark/DCGC-19255/2020\|EPI_ISL_748300\|202 | ···················· | ···················· | ····················· | ···················· | ···················· | ···················· | |
| AA | 514 hCoV-19/Denmark/DCGC-19287/2020\|EPI_ISL_748301\|202 | ···················· | ···················· | ····················· | ···················· | ···················· | ···················· | |
| AA | 515 hCoV-19/Denmark/DCGC-19297/2020\|EPI_ISL_748302\|202 | ···················· | ···················· | ····················· | ···················· | ···················· | ···················· | |
| AA | 516 hCoV-19/Denmark/DCGC-19369/2020\|EPI_ISL_748303\|202 | ···················· | ···················· | ····················· | ···················· | ···················· | ···················· | |
| AA | 517 hCoV-19/Denmark/DCGC-19378/2020\|EPI_ISL_748304\|202 | ···················· | ···················· | ····················· | ···················· | ···················· | ···················· | |
| AA | 518 hCoV-19/Denmark/DCGC-19397/2020\|EPI_ISL_748305\|202 | ···················· | ···················· | ····················· | ···················· | ···················· | ···················· | |
| AA | 519 hCoV-19/Denmark/DCGC-19405/2020\|EPI_ISL_748306\|202 | ···················· | ···················· | ····················· | ···················· | ···················· | ···················· | |
| AA | 520 hCoV-19/Denmark/DCGC-19409/2020\|EPI_ISL_748307\|202 | ···················· | ···················· | ····················· | ···················· | ···················· | ···················· | |
| AA | 521 hCoV-19/Denmark/DCGC-19415/2020\|EPI_ISL_748308\|202 | ···················· | ···················· | ····················· | ···················· | ···················· | ···················· | |
| AA | 522 hCoV-19/Denmark/DCGC-19421/2020\|EPI_ISL_748309\|202 | ···················· | ···················· | ····················· | ···················· | ···················· | ···················· | |
| AA | 523 hCoV-19/Denmark/DCGC-17852/2020\|EPI_ISL_749600\|202 | ···················· | ···················· | ····················· | ···················· | ···················· | ···················· | |
| AA | 524 hCoV-19/Denmark/DCGC-17855/2020\|EPI_ISL_749601\|202 | ···················· | ··········A········· | ····················· | ···················· | ···················· | ···················· | |
| AA | 525 hCoV-19/Denmark/DCGC-17857/2020\|EPI_ISL_749602\|202 | ···················· | ···················· | ····················· | ···················· | ···················· | ···················· | |
| AA | 526 hCoV-19/Denmark/DCGC-17859/2020\|EPI_ISL_749603\|202 | ···················· | ···················· | ····················· | ···················· | ···················· | ···················· | |
| AA | 527 hCoV-19/Denmark/DCGC-17860/2020\|EPI_ISL_749604\|202 | ···················· | ···················· | ····················· | ···················· | ···················· | ···················· | |
| AA | 528 hCoV-19/Denmark/DCGC-17861/2020\|EPI_ISL_749605\|202 | ···················· | ···················· | ····················· | ···················· | ···················· | ···················· | |
| AA | 529 hCoV-19/Denmark/DCGC-17863/2020\|EPI_ISL_749606\|202 | ···················· | ···················· | ····················· | ···················· | ···················· | ···················· | |
| AA | 530 hCoV-19/Denmark/DCGC-17864/2020\|EPI_ISL_749607\|202 | ···················· | ···················· | ····················· | ···················· | ···················· | ···················· | |
| AA | 531 hCoV-19/Denmark/DCGC-17866/2020\|EPI_ISL_749608\|202 | ···················· | ···················· | ····················· | ···················· | ···················· | ···················· | |
| AA | 532 hCoV-19/Denmark/DCGC-17867/2020\|EPI_ISL_749609\|202 | ···················· | ···················· | ····················· | ···················· | ···················· | ···················· | |
| AA | 533 hCoV-19/Denmark/DCGC-17868/2020\|EPI_ISL_749610\|202 | ···················· | ·A·················· | ····················· | ···················· | ···················· | ···················· | |
| AA | 534 hCoV-19/Denmark/DCGC-17869/2020\|EPI_ISL_749611\|202 | ···················· | ···················· | ····················· | ···················· | ···················· | ···················· | |
| AA | 535 hCoV-19/Denmark/DCGC-17871/2020\|EPI_ISL_749612\|202 | ···················· | ···················· | ····················· | ···················· | ···················· | ···················· | |
| AA | 536 hCoV-19/Denmark/DCGC-17873/2020\|EPI_ISL_749613\|202 | ···················· | ···················· | ····················· | ···················· | ···················· | ···················· | |
| AA | 537 hCoV-19/Denmark/DCGC-17874/2020\|EPI_ISL_749614\|202 | ···················· | ·A·················· | ····················· | ···················· | ···················· | ···················· | |
| AA | 538 hCoV-19/Denmark/DCGC-17875/2020\|EPI_ISL_749615\|202 | ···················· | ···················· | ····················· | ···················· | ···················· | ···················· | |
| AA | 539 hCoV-19/Denmark/DCGC-17877/2020\|EPI_ISL_749616\|202 | ···················· | ···················· | ····················· | ···················· | ···················· | ···················· | |
| AA | 540 hCoV-19/Denmark/DCGC-17878/2020\|EPI_ISL_749617\|202 | ···················· | ···················· | ····················· | ···················· | ···················· | ···················· | |
| AA | 541 hCoV-19/Denmark/DCGC-17880/2020\|EPI_ISL_749618\|202 | ···················· | ···················· | ····················· | ···················· | ···················· | ···················· | |
| AA | 542 hCoV-19/Denmark/DCGC-17881/2020\|EPI_ISL_749619\|202 | ···················· | ···················· | ····················· | ···················· | ···················· | ···················· | |
| AA | 543 hCoV-19/Denmark/DCGC-17882/2020\|EPI_ISL_749620\|202 | ···················· | ···················· | ····················· | ···················· | ···················· | ···················· | |
| AA | 544 hCoV-19/Denmark/DCGC-17883/2020\|EPI_ISL_749621\|202 | ···················· | ···················· | ····················· | ···················· | ···················· | ···················· | |
| AA | 545 hCoV-19/Denmark/DCGC-17885/2020\|EPI_ISL_749622\|202 | ···················· | ···················· | ····················· | ···················· | ···················· | ···················· | |
| AA | 546 hCoV-19/Denmark/DCGC-17886/2020\|EPI_ISL_749623\|202 | ···················· | ···················· | ····················· | ···················· | ···················· | ···················· | |
| AA | 547 hCoV-19/Denmark/DCGC-17888/2020\|EPI_ISL_749624\|202 | ···················· | ·A·················· | ····················· | ···················· | ···················· | ···A················ | |
| AA | 548 hCoV-19/Denmark/DCGC-17889/2020\|EPI_ISL_749625\|202 | ···················· | ···················· | ····················· | ···················· | ···················· | ···················· | |
| AA | 549 hCoV-19/Denmark/DCGC-17891/2020\|EPI_ISL_749626\|202 | ···················· | ···················· | ····················· | ···················· | ···················· | ···················· | |
| AA | 550 hCoV-19/Denmark/DCGC-17893/2020\|EPI_ISL_749627\|202 | ···················· | ···················· | ····················· | ···················· | ···················· | ···················· | |
| AA | 551 hCoV-19/Denmark/DCGC-17894/2020\|EPI_ISL_749628\|202 | ···················· | ···················· | ····················· | ···················· | ···················· | ···················· | |
| AA | 552 hCoV-19/Denmark/DCGC-17895/2020\|EPI_ISL_749629\|202 | ···················· | ···················· | ····················· | ···················· | ···················· | ···················· | |
| AA | 553 hCoV-19/Denmark/DCGC-17897/2020\|EPI_ISL_749630\|202 | ···················· | ···················· | ····················· | ···················· | ···················· | ···················· | |
| AA | 554 hCoV-19/Denmark/DCGC-17898/2020\|EPI_ISL_749631\|202 | ···················· | ···················· | ····················· | ···················· | ···················· | ···················· | |
| AA | 555 hCoV-19/Denmark/DCGC-17899/2020\|EPI_ISL_749632\|202 | ···················· | ···················· | ····················· | ···················· | ···················· | ···················· | |
| AA | 556 hCoV-19/Denmark/DCGC-17901/2020\|EPI_ISL_749633\|202 | ···················· | ···················· | ····················· | ···················· | ···················· | ···················· | |

```
AA 557 hCoV-19/Denmark/DCGC-17903/2020|EPI_ISL_749634|202 ···················· ···················· ···················· ···················· ···················· ····················
AA 558 hCoV-19/Denmark/DCGC-17904/2020|EPI_ISL_749635|202 ···················· ···················· ···················· ···················· ···················· ····················
AA 559 hCoV-19/Denmark/DCGC-17905/2020|EPI_ISL_749636|202 ···················· ···················· ···················· ···················· ···················· ····················
AA 560 hCoV-19/Denmark/DCGC-17906/2020|EPI_ISL_749637|202 ···················· ···················· ···················· ···················· ···················· ····················
AA 561 hCoV-19/Denmark/DCGC-17909/2020|EPI_ISL_749638|202 ···················· ···················· ···················· ···················· ···················· ····················
AA 562 hCoV-19/Denmark/DCGC-17911/2020|EPI_ISL_749639|202 ···················· ···················· ···················· ···················· ···················· ····················
AA 563 hCoV-19/Denmark/DCGC-17974/2020|EPI_ISL_749673|202 ···················· ···················· ···················· ···················· ···················· ····················
AA 564 hCoV-19/Denmark/DCGC-17976/2020|EPI_ISL_749674|202 ···················· ···················· ···················· ···················· ···················· ····················
AA 565 hCoV-19/Denmark/DCGC-17977/2020|EPI_ISL_749675|202 ···················· ···················· ···················· ···················· ···················· ····················
AA 566 hCoV-19/Denmark/DCGC-17978/2020|EPI_ISL_749676|202 ···················· ···················· ···················· ···················· ···················· ····················
AA 567 hCoV-19/Denmark/DCGC-17980/2020|EPI_ISL_749677|202 ···················· ···················· ···················· ···················· ···················· ····················
AA 568 hCoV-19/Denmark/DCGC-17981/2020|EPI_ISL_749678|202 ···················· ···················· ···················· ···················· ···················· ····················
AA 569 hCoV-19/Denmark/DCGC-17982/2020|EPI_ISL_749679|202 ···················· ·············A······ ···················· ···················· ···················· ····················
AA 570 hCoV-19/Denmark/DCGC-16850/2020|EPI_ISL_750600|202 ···················· ···················· ···················· ···················· ···················· ····················
AA 571 hCoV-19/Denmark/DCGC-16853/2020|EPI_ISL_750601|202 ···················· ···················· ···················· ···················· ···················· ····················
AA 572 hCoV-19/Denmark/DCGC-16856/2020|EPI_ISL_750602|202 ···················· ···················· ···················· ···················· ···················· ····················
AA 573 hCoV-19/Denmark/DCGC-16857/2020|EPI_ISL_750603|202 ···················· ···················· ···················· ···················· ···················· ····················
AA 574 hCoV-19/Denmark/DCGC-16859/2020|EPI_ISL_750604|202 ···················· ···················· ···················· ···················· ···················· ····················
AA 575 hCoV-19/Denmark/DCGC-16860/2020|EPI_ISL_750605|202 ···················· ···················· ···················· ···················· ···················· ····················
AA 576 hCoV-19/Denmark/DCGC-16861/2020|EPI_ISL_750606|202 ···················· ···················· ···················· ···················· ···················· ····················
AA 577 hCoV-19/Denmark/DCGC-16862/2020|EPI_ISL_750607|202 ···················· ···················· ···················· ···················· ···················· ····················
AA 578 hCoV-19/Denmark/DCGC-16863/2020|EPI_ISL_750608|202 ···················· ···················· ···················· ···················· ···················· ····················
AA 579 hCoV-19/Denmark/DCGC-16865/2020|EPI_ISL_750609|202 ···················· ···················· ···················· ···················· ···················· ····················
AA 580 hCoV-19/Denmark/DCGC-16866/2020|EPI_ISL_750610|202 ···················· ···················· ···················· ···················· ···················· ····················
AA 581 hCoV-19/Denmark/DCGC-16868/2020|EPI_ISL_750611|202 ···················· ···················· ···················· ···················· ···················· ····················
AA 582 hCoV-19/Denmark/DCGC-16869/2020|EPI_ISL_750612|202 ···················· ···················· ···················· ···················· ···················· ····················
AA 583 hCoV-19/Denmark/DCGC-16874/2020|EPI_ISL_750613|202 ···················· ···················· ···················· ···················· ···················· ····················
AA 584 hCoV-19/Denmark/DCGC-16875/2020|EPI_ISL_750614|202 ···················· ···················· ···················· ···················· ···················· ····················
AA 585 hCoV-19/Denmark/DCGC-16876/2020|EPI_ISL_750615|202 ···················· ···················· ···················· ···················· ···················· ····················
AA 586 hCoV-19/Denmark/DCGC-16877/2020|EPI_ISL_750616|202 ···················· ···················· ···················· ···················· ···················· ····················
AA 587 hCoV-19/Denmark/DCGC-16878/2020|EPI_ISL_750617|202 ···················· ···················· ···················· ···················· ···················· ····················
AA 588 hCoV-19/Denmark/DCGC-16879/2020|EPI_ISL_750618|202 ···················· ···················· ···················· ···················· ···················· ····················
AA 589 hCoV-19/Denmark/DCGC-16881/2020|EPI_ISL_750619|202 ···················· ···················· ···················· ···················· ···················· ····················
AA 590 hCoV-19/Denmark/DCGC-16954/2020|EPI_ISL_750664|202 ···················· ···················· ···················· ···················· ···················· ····················
AA 591 hCoV-19/Denmark/DCGC-16956/2020|EPI_ISL_750665|202 ···················· ···················· ···················· ···················· ···················· ····················
AA 592 hCoV-19/Denmark/DCGC-16957/2020|EPI_ISL_750666|202 ···················· ···················· ···················· ···················· ···················· ····················
AA 593 hCoV-19/Denmark/DCGC-16959/2020|EPI_ISL_750667|202 ···················· ···················· ···················· ···················· ···················· ····················
AA 594 hCoV-19/Denmark/DCGC-16961/2020|EPI_ISL_750668|202 ···················· ···················· ···················· ···················· ···················· ····················
AA 595 hCoV-19/Denmark/DCGC-16962/2020|EPI_ISL_750669|202 ···················· ···················· ···················· ···················· ···················· ····················
AA 596 hCoV-19/Singapore/1491/2020|EPI_ISL_754100|2020-12 ···················· ···················· ···················· ···················· ···················· ···A················
AA 597 hCoV-19/Singapore/1492/2020|EPI_ISL_754101|2020-12 ···················· ···················· ···················· ···················· ···················· ···A················
AA 598 hCoV-19/Singapore/1493/2020|EPI_ISL_754102|2020-12 ···················· ···················· ···················· ···················· ···················· ···A················
AA 599 hCoV-19/Singapore/1494/2020|EPI_ISL_754103|2020-12 ···················· ···················· ···················· ···················· ···················· ···A················
AA 600 hCoV-19/Singapore/1495/2020|EPI_ISL_754104|2020-12 ···················· ···················· ···················· ···················· ···················· ····················
AA 601 hCoV-19/Singapore/1496/2020|EPI_ISL_754105|2020-12 ···················· ···················· ···················· ···················· ···················· ····················
AA 602 hCoV-19/Singapore/1497/2020|EPI_ISL_754106|2020-12 ···················· ···················· ···················· ···················· ···················· ····················
AA 603 hCoV-19/USA/CA-LACPHL-AF00035/2020|EPI_ISL_754118| ···················· ···················· ···················· ···················· ···················· ····················
AA 604 hCoV-19/USA/CA-LACPHL-AF00036/2020|EPI_ISL_754119| ···················· ···················· ···················· ···················· ···················· ····················
AA 605 hCoV-19/USA/CA-LACPHL-AF00037/2020|EPI_ISL_754120| ···················· ···················· ···················· ···················· ···················· ····················
AA 606 hCoV-19/USA/CA-LACPHL-AF00038/2020|EPI_ISL_754121| ···················· ·A·················· ···················· ···················· ···················· ····················
AA 608 hCoV-19/USA/CA-LACPHL-AF00047/2020|EPI_ISL_754123| ···················· ···················· ···················· ···················· ···················· ····················
AA 609 hCoV-19/USA/CA-LACPHL-AF00049/2020|EPI_ISL_754124| ···················· ···················· ···················· ···················· ···················· ····················
AA 610 hCoV-19/USA/CA-LACPHL-AF00050/2020|EPI_ISL_754125| ···················· ···················· ···················· ···················· ···················· ····················
AA 611 hCoV-19/USA/CA-LACPHL-AF00051/2020|EPI_ISL_754126| ···················· ···················· ···················· ···················· ···················· ····················
AA 612 hCoV-19/USA/CA-LACPHL-AF00052/2020|EPI_ISL_754127| ···················· ···················· ···················· ···················· ···················· ····················
AA 613 hCoV-19/USA/CA-LACPHL-AF00053/2020|EPI_ISL_754128| ···················· ···················· ···················· ···················· ···················· ····················
AA 614 hCoV-19/USA/CA-LACPHL-AF00054/2020|EPI_ISL_754129| ···················· ···················· ···················· ···················· ···················· ····················
AA 615 hCoV-19/USA/CA-LACPHL-AF00055/2020|EPI_ISL_754130| ···················· ···················· ···················· ···················· ···················· ····················
AA 616 hCoV-19/USA/CA-LACPHL-AF00056/2020|EPI_ISL_754131| ···················· ···················· ···················· ···················· ···················· ····················
AA 617 hCoV-19/USA/CA-LACPHL-AF00057/2020|EPI_ISL_754132| ···················· ···················· ···················· ···················· ···················· ····················
AA 618 hCoV-19/USA/CA-LACPHL-AF00058/2020|EPI_ISL_754133| ···················· ···················· ···················· ···················· ···················· ····················
AA 619 hCoV-19/USA/CA-LACPHL-AF00059/2020|EPI_ISL_754134| ···················· ·A·················· ···················· ···················· ···················· ····················
AA 620 hCoV-19/USA/CA-LACPHL-AF00060/2020|EPI_ISL_754135| ···················· ·A·················· ···················· ···················· ···················· ····················
AA 621 hCoV-19/France/OCC-182/2020|EPI_ISL_754136|2020-09 ···················· ···················· ···················· ···················· ···················· ····················
AA 622 hCoV-19/France/OCC-183/2020|EPI_ISL_754137|2020-10 ···················· ···················· ···················· ···················· ···················· ····················
AA 623 hCoV-19/France/OCC-184/2020|EPI_ISL_754138|2020-10 ···················· ···················· ···················· ···················· ···················· ····················
AA 624 hCoV-19/France/OCC-185/2020|EPI_ISL_754139|2020-11 ···················· ···················· ···················· ···················· ···················· ····················
AA 625 hCoV-19/France/BRE-IPP10826/2020|EPI_ISL_754141|20 ···················· ···················· ···················· ···················· ···················· ····················
AA 626 hCoV-19/France/BRE-IPP10827/2020|EPI_ISL_754142|20 ···················· ···················· ···················· ···················· ···················· ····················
AA 627 hCoV-19/France/BRE-IPP10828/2020|EPI_ISL_754143|20 ···················· ···················· ···················· ···················· ···················· ····················
AA 628 hCoV-19/France/BRE-IPP10831/2020|EPI_ISL_754144|20 ···················· ···················· ···················· ···················· ···················· ····················
AA 629 hCoV-19/France/BRE-IPP10838/2020|EPI_ISL_754145|20 ···················· ···················· ···················· ···················· ···················· ····················
AA 630 hCoV-19/France/BRE-IPP10855/2020|EPI_ISL_754147|20 ···················· ···················· ···················· ···················· ···················· ····················
AA 631 hCoV-19/France/BRE-IPP10856/2020|EPI_ISL_754148|20 ···················· ···················· ···················· ···················· ···················· ····················
AA 632 hCoV-19/France/BRE-IPP10871/2020|EPI_ISL_754149|20 ···················· ···················· ···················· ···················· ···················· ····················
AA 633 hCoV-19/France/BRE-IPP10873/2020|EPI_ISL_754150|20 ···················· ···················· ···················· ···················· ···················· ····················
AA 634 hCoV-19/France/BRE-IPP10879/2020|EPI_ISL_754151|20 ···················· ···················· ···················· ···················· ···················· ····················
AA 635 hCoV-19/France/BRE-IPP10833/2020|EPI_ISL_754152|20 ···················· ···················· ···················· ···················· ···················· ····················
AA 636 hCoV-19/France/BRE-IPP10835/2020|EPI_ISL_754153|20 ···················· ···················· ···················· ···················· ···················· ····················
AA 637 hCoV-19/France/BRE-IPP10841/2020|EPI_ISL_754154|20 ···················· ···················· ···················· ···················· ···················· ····················
```

| | | | | | | | | |
|---|---|---|---|---|---|---|---|---|
| AA | 638 hCoV-19/France/BRE-IPP10843/2020\|EPI_ISL_754155\|20 | ···················· | ···················· | ···················· | ···················· | ···················· | ···················· | |
| AA | 639 hCoV-19/France/BRE-IPP10844/2020\|EPI_ISL_754156\|20 | ···················· | ···················· | ···················· | ···················· | ···················· | ···················· | |
| AA | 640 hCoV-19/France/BRE-IPP10853/2020\|EPI_ISL_754157\|20 | ···················· | ···················· | ···················· | ···················· | ···················· | ···················· | |
| AA | 641 hCoV-19/France/BRE-IPP10858/2020\|EPI_ISL_754158\|20 | ···················· | ···················· | ···················· | ···················· | ···················· | ···················· | |
| AA | 642 hCoV-19/France/BRE-IPP10863/2020\|EPI_ISL_754159\|20 | ···················· | ···················· | ···················· | ···················· | ···················· | ···················· | |
| AA | 643 hCoV-19/France/BRE-IPP10864/2020\|EPI_ISL_754160\|20 | ···················· | ···················· | ···················· | ···················· | ···················· | ···················· | |
| AA | 644 hCoV-19/France/BRE-IPP10865/2020\|EPI_ISL_754161\|20 | ···················· | ···················· | ···················· | ···················· | ···················· | ···················· | |
| AA | 645 hCoV-19/France/BRE-IPP10866/2020\|EPI_ISL_754162\|20 | ···················· | ···················· | ···················· | ···················· | ···················· | ···················· | |
| AA | 646 hCoV-19/France/BRE-IPP10867/2020\|EPI_ISL_754163\|20 | ···················· | ···················· | ···················· | ···················· | ···················· | ···················· | |
| AA | 647 hCoV-19/France/BRE-IPP10868/2020\|EPI_ISL_754164\|20 | ···················· | ···················· | ···················· | ···················· | ···················· | ···················· | |
| AA | 648 hCoV-19/France/BRE-IPP10872/2020\|EPI_ISL_754165\|20 | ···················· | ···················· | ···················· | ···················· | ···················· | ···················· | |
| AA | 649 hCoV-19/France/BRE-IPP10874/2020\|EPI_ISL_754166\|20 | ···················· | ···················· | ···················· | ···················· | ···················· | ···················· | |
| AA | 650 hCoV-19/France/BRE-IPP10877/2020\|EPI_ISL_754167\|20 | ···················· | ···················· | ···················· | ···················· | ···················· | ···················· | |
| AA | 651 hCoV-19/France/BRE-IPP10878/2020\|EPI_ISL_754168\|20 | ···················· | ···················· | ···················· | ···················· | ···················· | ···················· | |
| AA | 652 hCoV-19/France/BRE-IPP10836/2020\|EPI_ISL_754169\|20 | ···················· | ···················· | ···················· | ···················· | ···················· | ···················· | |
| AA | 653 hCoV-19/France/BRE-IPP10875/2020\|EPI_ISL_754170\|20 | ···················· | ···················· | ···················· | ···················· | ···················· | ···················· | |
| AA | 654 hCoV-19/France/BRE-IPP10876/2020\|EPI_ISL_754171\|20 | ···················· | ···················· | ···················· | ···················· | ···················· | ···················· | |
| AA | 655 hCoV-19/France/BRE-IPP10881/2020\|EPI_ISL_754172\|20 | ···················· | ···················· | ···················· | ···················· | ···················· | ···················· | |
| AA | 656 hCoV-19/Bangladesh/SUST-GEB-Habiganj-3/2020\|EPI_IS | ···················· | ···················· | ···················· | ···················· | ···················· | ···················· | |
| AA | 657 hCoV-19/Germany/DE-NI-ChVir21542/2020\|EPI_ISL_7541 | ···················· | ···················· | ···················· | ···················· | ······T············· | ···················· | |
| AA | 658 hCoV-19/Bangladesh/SUST-GEB-Sunamganj-5/2020\|EPI_I | ···················· | ···················· | ···················· | ···················· | ···················· | ···················· | |
| AA | 659 hCoV-19/Bangladesh/SUST-GEB-Habiganj-4/2020\|EPI_IS | ···················· | ···················· | ···················· | ···················· | ···················· | ···················· | |
| AA | 660 hCoV-19/Bangladesh/SUST-GEB-Habiganj-6/2020\|EPI_IS | ···················· | ···················· | ···················· | ···················· | ···················· | ···················· | |
| AA | 661 hCoV-19/Georgia/Tb-86952/2020\|EPI_ISL_754180\|2020- | ···················· | ···················· | ···················· | ···················· | ···················· | ···················· | |
| AA | 662 hCoV-19/Georgia/Tb-97937/2020\|EPI_ISL_754181\|2020- | ···················· | ···················· | ···················· | ···················· | ···················· | ···················· | |
| AA | 663 hCoV-19/USA/WY-WYPHL-20092968/2020\|EPI_ISL_754182\| | ···················· | ···················· | ···················· | ···················· | ···················· | ···················· | |
| AA | 664 hCoV-19/USA/WY-WYPHL-20094896/2020\|EPI_ISL_754183\| | ···················· | ···················· | ···················· | ···················· | ···················· | ···················· | |
| AA | 665 hCoV-19/Germany/BE-ChVir-W2121-4431/2020\|EPI_ISL_7 | ···················· | ·A·················· | ···················· | ···················· | ···················· | ··A················· | |
| AA | 666 hCoV-19/Germany/BE-ChVir-D1981-5990/2020\|EPI_ISL_7 | ···················· | ·A·················· | ···················· | ···················· | ···················· | ··A················· | |
| AA | 667 hCoV-19/Germany/BE-ChVir-D1917-8291/2020\|EPI_ISL_7 | ···················· | ·A·················· | ···················· | ···················· | ···················· | ··A················· | |
| AA | 668 hCoV-19/Germany/BE-ChVir-D1947-5915/2020\|EPI_ISL_7 | ···················· | ·A·················· | ···················· | ···················· | ···················· | ··A················· | |
| AA | 669 hCoV-19/Germany/BE-ChVir-W2120-5976/2020\|EPI_ISL_7 | ···················· | ·A·················· | ···················· | ···················· | ···················· | ··A················· | |
| AA | 670 hCoV-19/Germany/BE-ChVir-D2028-8264/2020\|EPI_ISL_7 | ···················· | ···················· | ···················· | ···················· | ···················· | ···················· | |
| AA | 671 hCoV-19/Germany/BE-ChVir-W2127-8203/2020\|EPI_ISL_7 | ···················· | ···················· | ···················· | ···················· | ···················· | ···················· | |
| AA | 672 hCoV-19/Germany/NW-ChVir-D1925-5947/2020\|EPI_ISL_7 | ···················· | ·A·················· | ···················· | ···················· | ···················· | ··A················· | |
| AA | 673 hCoV-19/Germany/BE-ChVir-D2019-4498/2020\|EPI_ISL_7 | ···················· | ···················· | ···················· | ···················· | ···················· | ···················· | |
| AA | 674 hCoV-19/Russia/MOS-CRIE-13606547/2020\|EPI_ISL_7541 | ···················· | ···················· | ···················· | ···················· | ···················· | ···················· | |
| AA | 675 hCoV-19/Russia/MOS-CRIE-13604992/2020\|EPI_ISL_7541 | ···················· | ···················· | ···················· | ···················· | ···················· | ···················· | |
| AA | 676 hCoV-19/Russia/MOS-CRIE-13604226/2020\|EPI_ISL_7541 | ···················· | ···················· | ···················· | ···················· | ···················· | ···················· | |
| AA | 677 hCoV-19/Russia/MOS-CRIE-13642264/2020\|EPI_ISL_7541 | ············A······· | ···················· | ···················· | ···················· | ···················· | ···················· | |
| AA | 678 hCoV-19/USA/ME-HETL-J0244/2020\|EPI_ISL_755400\|2020 | ···················· | ···················· | ···················· | ···················· | ···················· | ···················· | |
| AA | 679 hCoV-19/USA/ME-HETL-J0246/2020\|EPI_ISL_755401\|2020 | ···················· | ·A·················· | ···················· | ···················· | ···················· | ···················· | |
| AA | 680 hCoV-19/USA/ME-HETL-J0247/2020\|EPI_ISL_755402\|2020 | ···················· | ···················· | ···················· | ···················· | ···················· | ···················· | |
| AA | 681 hCoV-19/USA/ME-HETL-J0250/2020\|EPI_ISL_755403\|2020 | ···················· | ···················· | ···················· | ···················· | ···················· | ···················· | |
| AA | 682 hCoV-19/USA/ME-HETL-J0251/2020\|EPI_ISL_755404\|2020 | ···················· | ···················· | ···················· | ···················· | ···················· | ···················· | |
| AA | 683 hCoV-19/USA/ME-HETL-J0256/2020\|EPI_ISL_755405\|2020 | ···················· | ···················· | ···················· | ···················· | ···················· | ···················· | |
| AA | 684 hCoV-19/USA/ME-HETL-J0258/2020\|EPI_ISL_755406\|2020 | ···················· | ···················· | ···················· | ···················· | ···················· | ···················· | |
| AA | 685 hCoV-19/USA/ME-HETL-J0259/2020\|EPI_ISL_755407\|2020 | ···················· | ···················· | ···················· | ···················· | ···················· | ···················· | |
| AA | 686 hCoV-19/USA/ME-HETL-J0260/2020\|EPI_ISL_755408\|2020 | ···················· | ···················· | ···················· | ···················· | ···················· | ···················· | |
| AA | 687 hCoV-19/USA/ME-HETL-J0263/2020\|EPI_ISL_755410\|2020 | ···················· | ···················· | ···················· | ···················· | ···················· | ···················· | |
| AA | 688 hCoV-19/USA/ME-HETL-J0264/2020\|EPI_ISL_755411\|2020 | ···················· | ···················· | ···················· | ···················· | ···················· | ···················· | |
| AA | 689 hCoV-19/USA/ME-HETL-J0266/2020\|EPI_ISL_755412\|2020 | ···················· | ···················· | ···················· | ···················· | ···················· | ···················· | |
| AA | 690 hCoV-19/USA/ME-HETL-J0268/2020\|EPI_ISL_755414\|2020 | ···················· | ·A·················· | ···················· | ···················· | ···················· | ···················· | |
| AA | 691 hCoV-19/USA/ME-HETL-J0270/2020\|EPI_ISL_755416\|2020 | ···················· | ···················· | ···················· | ···················· | ···················· | ···················· | |
| AA | 692 hCoV-19/USA/ME-HETL-J0272/2020\|EPI_ISL_755418\|2020 | ···················· | ···················· | ···················· | ···················· | ···················· | ···················· | |
| AA | 693 hCoV-19/USA/ME-HETL-J0273/2020\|EPI_ISL_755419\|2020 | ··················T· | ···················· | ···················· | ···················· | ···················· | ···················· | 1 |
| AA | 694 hCoV-19/USA/ME-HETL-J0275/2020\|EPI_ISL_755421\|2020 | ···················· | ···················· | ···················· | ···················· | ···················· | ···················· | |
| AA | 695 hCoV-19/USA/ME-HETL-J0276/2020\|EPI_ISL_755422\|2020 | ···················· | ···················· | ···················· | ···················· | ···················· | ···················· | |
| AA | 696 hCoV-19/USA/ME-HETL-J0277/2020\|EPI_ISL_755423\|2020 | ···················· | ···················· | ···················· | ···················· | ···················· | ···················· | |
| AA | 697 hCoV-19/USA/ME-HETL-J0278/2020\|EPI_ISL_755424\|2020 | ···················· | ···················· | ···················· | ···················· | ···················· | ···················· | |
| AA | 698 hCoV-19/USA/ME-HETL-J0279/2020\|EPI_ISL_755425\|2020 | ···················· | ···················· | ···················· | ···················· | ···················· | ···················· | |
| AA | 699 hCoV-19/USA/ME-HETL-J0284/2020\|EPI_ISL_755427\|2020 | ···················· | ···················· | ···················· | ···················· | ···················· | ···················· | |
| AA | 700 hCoV-19/USA/ME-HETL-J0285/2020\|EPI_ISL_755428\|2020 | ···················· | ·A·················· | ·········T·········· | ···················· | ···················· | ···················· | |
| AA | 701 hCoV-19/USA/ME-HETL-J0286/2020\|EPI_ISL_755429\|2020 | ···················· | ·A·················· | ·········T·········· | ···················· | ···················· | ···················· | |
| AA | 702 hCoV-19/USA/ME-HETL-J0287/2020\|EPI_ISL_755430\|2020 | ···················· | ···················· | ···················· | ···················· | ···················· | ···················· | |
| AA | 703 hCoV-19/USA/ME-HETL-J0288/2020\|EPI_ISL_755431\|2020 | ···················· | ···················· | ···················· | ···················· | ···················· | ···················· | |
| AA | 704 hCoV-19/USA/ME-HETL-J0290/2020\|EPI_ISL_755432\|2020 | ···················· | ···················· | ···················· | ···················· | ···················· | ···················· | |
| AA | 705 hCoV-19/USA/ME-HETL-J0291/2020\|EPI_ISL_755433\|2020 | ···················· | ···················· | ···················· | ···················· | ···················· | ···················· | |
| AA | 706 hCoV-19/USA/ME-HETL-J0294/2020\|EPI_ISL_755434\|2020 | ···················· | ···················· | ···················· | ···················· | ···················· | ···················· | |
| AA | 707 hCoV-19/USA/ME-HETL-J0298/2020\|EPI_ISL_755436\|2020 | ···················· | ···················· | ···················· | ···················· | ···················· | ···················· | |
| AA | 708 hCoV-19/USA/ME-HETL-J0299/2020\|EPI_ISL_755437\|2020 | ···················· | ·A·················· | ···················· | ···················· | ···················· | ···················· | |
| AA | 709 hCoV-19/USA/ME-HETL-J0302/2020\|EPI_ISL_755440\|2020 | ···················· | ·A·················· | ···················· | ···················· | ···················· | ···················· | |
| AA | 710 hCoV-19/USA/ME-HETL-J0320/2020\|EPI_ISL_755449\|2020 | ···················· | ···················· | ···················· | ···················· | ···················· | ···················· | |
| AA | 711 hCoV-19/USA/ME-HETL-J0321/2020\|EPI_ISL_755450\|2020 | ···················· | ···················· | ···················· | ···················· | ···················· | ···················· | |
| AA | 712 hCoV-19/USA/ME-HETL-J0322/2020\|EPI_ISL_755451\|2020 | ···················· | ···················· | ···················· | ···················· | ···················· | ···················· | |
| AA | 713 hCoV-19/USA/ME-HETL-J0323/2020\|EPI_ISL_755452\|2020 | ···················· | ···················· | ···················· | ···················· | ···················· | ···················· | |
| AA | 715 hCoV-19/USA/ME-HETL-J0329/2020\|EPI_ISL_755457\|2020 | ···················· | ···················· | ···················· | ···················· | ···················· | ···················· | |
| AA | 716 hCoV-19/USA/ME-HETL-J0330/2020\|EPI_ISL_755458\|2020 | ···················· | ·············A······ | ···················· | ···················· | ···················· | ···················· | |
| AA | 717 hCoV-19/USA/ME-HETL-J0334/2020\|EPI_ISL_755462\|2020 | ···················· | ···················· | ···················· | ···················· | ···················· | ···················· | |
| AA | 718 hCoV-19/USA/ME-HETL-J0335/2020\|EPI_ISL_755463\|2020 | ···················· | ···················· | ···················· | ···················· | ···················· | ···················· | |

AA 719 hCoV-19/USA/ME-HETL-J0337/2020|EPI_ISL_755465|2020 .................... .................... .................... .................... .................... ....................
AA 721 hCoV-19/USA/ME-HETL-J0339/2020|EPI_ISL_755467|2020 .................... .................... .................... .................... .................... ....................
AA 722 hCoV-19/USA/ME-HETL-J0340/2020|EPI_ISL_755468|2020 .................... .................... .................... .................... .................... ....................
AA 723 hCoV-19/USA/ME-HETL-J0341/2020|EPI_ISL_755469|2020 .................... .................... .................... .................... .................... ....................
AA 724 hCoV-19/USA/ME-HETL-J0344/2020|EPI_ISL_755470|2020 .................... .................... .................... .................... .................... ....................
AA 725 hCoV-19/USA/ME-HETL-J0345/2020|EPI_ISL_755471|2020 .................... .................... .................... .................... .................... ....................
AA 726 hCoV-19/USA/ME-HETL-J0346/2020|EPI_ISL_755472|2020 .................... .................... .................... .................... .................... ....................
AA 727 hCoV-19/USA/ME-HETL-J0347/2020|EPI_ISL_755473|2020 .................... .................... .................... .................... .................... ....................
AA 728 hCoV-19/USA/ME-HETL-J0349/2020|EPI_ISL_755474|2020 .................... .................... .................... .................... .................... ....................
AA 729 hCoV-19/USA/ME-HETL-J0350/2020|EPI_ISL_755475|2020 .................... .................... .................... .................... .................... ....................
AA 730 hCoV-19/USA/ME-HETL-J0351/2020|EPI_ISL_755476|2020 .................... .................... .................... .................... .................... ....................
AA 731 hCoV-19/USA/ME-HETL-J0354/2020|EPI_ISL_755477|2020 .................... .................... .................... .................... .................... ....................
AA 732 hCoV-19/USA/ME-HETL-J0355/2020|EPI_ISL_755478|2020 .................... .................... .................... .................... .................... ....................
AA 733 hCoV-19/USA/ME-HETL-J0356/2020|EPI_ISL_755479|2020 .................... .................... .................... .................... .................... ....................
AA 734 hCoV-19/USA/ME-HETL-J0357/2020|EPI_ISL_755480|2020 .................... .................... .................... .................... .................... ....................
AA 735 hCoV-19/USA/ME-HETL-J0358/2020|EPI_ISL_755481|2020 .................... .A.................. .................... .................... .................... ....................
AA 736 hCoV-19/USA/ME-HETL-J0360/2020|EPI_ISL_755483|2020 .................... .................... .................... .................... .................... ....................
AA 737 hCoV-19/USA/ME-HETL-J0361/2020|EPI_ISL_755484|2020 .................... .................... .................... .................... .................... ....................
AA 738 hCoV-19/USA/ME-HETL-J0362/2020|EPI_ISL_755485|2020 .................... .................... .................... .................... .................... ....................
AA 739 hCoV-19/USA/ME-HETL-J0363/2020|EPI_ISL_755486|2020 .................... .................... .................... .................... .................... ....................
AA 740 hCoV-19/USA/ME-HETL-J0365/2020|EPI_ISL_755487|2020 .................... .................... .................... .................... .................... ....................
AA 741 hCoV-19/USA/ME-HETL-J0366/2020|EPI_ISL_755488|2020 .................... .A.................. .................... .................... .................... ....................
AA 742 hCoV-19/USA/ME-HETL-J0368/2020|EPI_ISL_755490|2020 .................... .................... .................... .................... .................... ....................
AA 743 hCoV-19/USA/ME-HETL-J0369/2020|EPI_ISL_755491|2020 .................... .A.................. .................... .................... .................... ....................
AA 744 hCoV-19/USA/ME-HETL-J0370/2020|EPI_ISL_755492|2020 .................... .A.................. .................... .................... .................... ....................
AA 745 hCoV-19/USA/ME-HETL-J0371/2020|EPI_ISL_755493|2020 .................... .A.................. .................... .................... .................... ....................
AA 746 hCoV-19/USA/ME-HETL-J0373/2020|EPI_ISL_755494|2020 .................... .................... .................... .................... .................... ....................
AA 747 hCoV-19/USA/ME-HETL-J0374/2020|EPI_ISL_755495|2020 .................... .A.................. .................... .................... .................... ....................
AA 748 hCoV-19/USA/ME-HETL-J0375/2020|EPI_ISL_755496|2020 .................... .................... .................... .................... .................... ....................
AA 749 hCoV-19/USA/ME-HETL-J0376/2020|EPI_ISL_755497|2020 .................... .................... .................... .................... .................... ....................
AA 750 hCoV-19/USA/ME-HETL-J0379/2020|EPI_ISL_755498|2020 .................... .A.................. .................... .................... .................... ....................
AA 751 hCoV-19/USA/ME-HETL-J0381/2020|EPI_ISL_755499|2020 .................... .................... .................... .................... .................... ....................
AA 752 hCoV-19/England/MILK-C7AB9B/2020|EPI_ISL_756725|20 .................... .................... .................... .................... .................... ....................
AA 753 hCoV-19/England/MILK-C7AE56/2020|EPI_ISL_756730|20 .................... .................... .................... .................... .................... ....................
AA 754 hCoV-19/England/QEUH-C7D864/2020|EPI_ISL_756737|20 .................... .................... .................... .................... .................... ....................
AA 755 hCoV-19/England/MILK-C784A1/2020|EPI_ISL_756741|20 .................... .................... .................... .................... .................... ....................
AA 756 hCoV-19/England/MILK-C7B23B/2020|EPI_ISL_756747|20 .................... .................... .................... .................... .................... ....................
AA 757 hCoV-19/England/QEUH-C7C838/2020|EPI_ISL_756748|20 .................... .................... .................... .................... .................... ....................
AA 758 hCoV-19/England/QEUH-C80084/2020|EPI_ISL_756750|20 .................... .................... .................... .................... ......T............. ....................
AA 759 hCoV-19/England/MILK-C78535/2020|EPI_ISL_756757|20 .................... .................... .................... .................... .................... ....................
AA 760 hCoV-19/England/QEUH-C7C7C2/2020|EPI_ISL_756761|20 .................... .................... .................... .................... .................... ....................
AA 761 hCoV-19/England/MILK-C7B5D8/2020|EPI_ISL_756762|20 .................... .................... .................... .................... .................... ....................
AA 762 hCoV-19/England/MILK-C7B73C/2020|EPI_ISL_756765|20 .................... .................... .................... .................... .................... ....................
AA 763 hCoV-19/England/MILK-C7B63F/2020|EPI_ISL_756770|20 .................... .................... .................... .................... .................... ....................
AA 764 hCoV-19/England/MILK-C7B59C/2020|EPI_ISL_756786|20 .................... .A.................. .................... .................... .................... ...A................
AA 765 hCoV-19/England/MILK-C7B523/2020|EPI_ISL_756787|20 .................... .................... .................... .................... .................... ....................
AA 766 hCoV-19/England/MILK-C782D4/2020|EPI_ISL_756791|20 .................... .................... .................... .................... .................... ....................
AA 767 hCoV-19/England/MILK-C7BA33/2020|EPI_ISL_756798|20 .................... .................... .................... .................... .................... ....................
AA 768 hCoV-19/England/MILK-C7B3B0/2020|EPI_ISL_756799|20 .................... .................... .................... .................... .................... ....................
AA 769 hCoV-19/Denmark/DCGC-19428/2020|EPI_ISL_748310|202 .................... .................... .................... .................... .................... ....................
AA 770 hCoV-19/Denmark/DCGC-19436/2020|EPI_ISL_748311|202 .................... .................... .................... .................... .................... ....................
AA 771 hCoV-19/Denmark/DCGC-19481/2020|EPI_ISL_748312|202 .................... .................... .................... .................... .................... ....................
AA 772 hCoV-19/Denmark/DCGC-19494/2020|EPI_ISL_748313|202 .................... .................... .................... .................... .................... ....................
AA 773 hCoV-19/Denmark/DCGC-19552/2020|EPI_ISL_748314|202 .................... .................... .................... .................... .................... ....................
AA 774 hCoV-19/Denmark/DCGC-19560/2020|EPI_ISL_748315|202 .................... .................... .................... .................... .................... ....................
AA 775 hCoV-19/Denmark/DCGC-19601/2020|EPI_ISL_748316|202 .................... .................... .................... .................... .................... ....................
AA 776 hCoV-19/Denmark/DCGC-19617/2020|EPI_ISL_748317|202 .................... .................... .................... .................... .................... ....................
AA 777 hCoV-19/Denmark/DCGC-19623/2020|EPI_ISL_748318|202 .................... .................... .................... .................... .................... ....................
AA 778 hCoV-19/Denmark/DCGC-19725/2020|EPI_ISL_748319|202 .................... .................... .................... .................... .................... ....................
AA 779 hCoV-19/Denmark/DCGC-19732/2020|EPI_ISL_748320|202 .................... .................... .................... .................... .................... ....................
AA 780 hCoV-19/Denmark/DCGC-19265/2020|EPI_ISL_748321|202 .................... .................... .................... .................... .................... ....................
AA 781 hCoV-19/Denmark/DCGC-19509/2020|EPI_ISL_748322|202 .................... .................... .................... .................... .................... ....................
AA 782 hCoV-19/Denmark/DCGC-18999/2020|EPI_ISL_748323|202 .................... .................... .................... .................... .................... ....................
AA 783 hCoV-19/Denmark/DCGC-19063/2020|EPI_ISL_748324|202 .................... .................... .................... .................... .................... ....................
AA 784 hCoV-19/Denmark/DCGC-18808/2020|EPI_ISL_748359|202 .................... .................... .................... .................... .................... ....................
AA 785 hCoV-19/Denmark/DCGC-18852/2020|EPI_ISL_748360|202 .................... .................... .................... .................... .................... ....................
AA 786 hCoV-19/Denmark/DCGC-18894/2020|EPI_ISL_748361|202 .................... .................... .................... .................... .................... ....................
AA 787 hCoV-19/Denmark/DCGC-18981/2020|EPI_ISL_748362|202 .................... .................... .................... .................... .................... ....................
AA 788 hCoV-19/Denmark/DCGC-19013/2020|EPI_ISL_748363|202 .................... .................... .................... .................... .................... ....................
AA 789 hCoV-19/Denmark/DCGC-19037/2020|EPI_ISL_748364|202 .................... .................... .................... .................... .................... ....................
AA 790 hCoV-19/Denmark/DCGC-19137/2020|EPI_ISL_748365|202 .................... .................... .................... .................... .................... ....................
AA 791 hCoV-19/Denmark/DCGC-19149/2020|EPI_ISL_748366|202 .................... .................... .................... .................... .................... ....................
AA 792 hCoV-19/Denmark/DCGC-19171/2020|EPI_ISL_748367|202 .................... .................... .................... .................... .................... ....................
AA 793 hCoV-19/Denmark/DCGC-19202/2020|EPI_ISL_748368|202 .................... .................... .................... .................... .................... ....................
AA 794 hCoV-19/Denmark/DCGC-19224/2020|EPI_ISL_748369|202 .................... .................... .................... .................... .................... ....................
AA 795 hCoV-19/Denmark/DCGC-19228/2020|EPI_ISL_748370|202 .................... .................... .................... .................... .................... ....................
AA 796 hCoV-19/Denmark/DCGC-19240/2020|EPI_ISL_748371|202 .................... .................... .................... .................... .................... ....................
AA 797 hCoV-19/Denmark/DCGC-19330/2020|EPI_ISL_748372|202 .................... .................... .................... .................... .................... ....................
AA 798 hCoV-19/Denmark/DCGC-19342/2020|EPI_ISL_748373|202 .................... .................... .................... .................... .................... ....................
AA 799 hCoV-19/Denmark/DCGC-19344/2020|EPI_ISL_748374|202 .................... .................... .................... .................... .................... ....................

AA 800 hCoV-19/Denmark/DCGC-19391/2020|EPI_ISL_748375|202 ···················· ···················· ···················· ···················· ···················· ····················
AA 801 hCoV-19/Denmark/DCGC-19496/2020|EPI_ISL_748376|202 ···················· ···················· ···················· ···················· ···················· ····················
AA 802 hCoV-19/Denmark/DCGC-19506/2020|EPI_ISL_748377|202 ···················· ···················· ···················· ···················· ···················· ····················
AA 803 hCoV-19/Denmark/DCGC-19520/2020|EPI_ISL_748378|202 ···················· ···················· ···················· ···················· ···················· ····················
AA 804 hCoV-19/Denmark/DCGC-19521/2020|EPI_ISL_748379|202 ···················· ···················· ···················· ···················· ···················· ····················
AA 805 hCoV-19/Denmark/DCGC-19540/2020|EPI_ISL_748380|202 ···················· ···················· ···················· ···················· ···················· ····················
AA 806 hCoV-19/Denmark/DCGC-19558/2020|EPI_ISL_748381|202 ···················· ···················· ···················· ···················· ···················· ····················
AA 807 hCoV-19/Denmark/DCGC-19592/2020|EPI_ISL_748382|202 ···················· ···················· ···················· ···················· ···················· ····················
AA 808 hCoV-19/Denmark/DCGC-19740/2020|EPI_ISL_748383|202 ···················· ···················· ···················· ···················· ···················· ····················
AA 809 hCoV-19/Denmark/DCGC-18756/2020|EPI_ISL_748384|202 ···················· ···················· ···················· ···················· ···················· ····················
AA 810 hCoV-19/Denmark/DCGC-18761/2020|EPI_ISL_748385|202 ···················· ···················· ···················· ···················· ···················· ····················
AA 811 hCoV-19/Denmark/DCGC-18778/2020|EPI_ISL_748386|202 ···················· ···················· ···················· ···················· ···················· ····················
AA 812 hCoV-19/Denmark/DCGC-18791/2020|EPI_ISL_748387|202 ···················· ···················· ···················· ···················· ···················· ····················
AA 813 hCoV-19/Denmark/DCGC-18792/2020|EPI_ISL_748388|202 ···················· ···················· ···················· ···················· ···················· ····················
AA 814 hCoV-19/Denmark/DCGC-18796/2020|EPI_ISL_748389|202 ···················· ···················· ···················· ···················· ···················· ····················
AA 815 hCoV-19/Denmark/DCGC-18823/2020|EPI_ISL_748390|202 ···················· ···················· ···················· ···················· ···················· ····················
AA 816 hCoV-19/Denmark/DCGC-18826/2020|EPI_ISL_748391|202 ···················· ···················· ···················· ···················· ···················· ····················
AA 817 hCoV-19/Denmark/DCGC-18830/2020|EPI_ISL_748392|202 ···················· ···················· ···················· ···················· ···················· ····················
AA 818 hCoV-19/Denmark/DCGC-18837/2020|EPI_ISL_748393|202 ···················· ···················· ···················· ···················· ···················· ····················
AA 819 hCoV-19/Denmark/DCGC-18840/2020|EPI_ISL_748394|202 ···················· ···················· ···················· ···················· ···················· ····················
AA 820 hCoV-19/Denmark/DCGC-18844/2020|EPI_ISL_748395|202 ···················· ···················· ···················· ···················· ···················· ····················
AA 821 hCoV-19/Denmark/DCGC-18869/2020|EPI_ISL_748396|202 ···················· ···················· ···················· ···················· ···················· ····················
AA 822 hCoV-19/Denmark/DCGC-18879/2020|EPI_ISL_748397|202 ···················· ···················· ···················· ···················· ···················· ····················
AA 823 hCoV-19/Denmark/DCGC-18882/2020|EPI_ISL_748398|202 ···················· ···················· ···················· ···················· ···················· ····················
AA 824 hCoV-19/Denmark/DCGC-18931/2020|EPI_ISL_748399|202 ···················· ···················· ···················· ···················· ··C················· ····················
AA 825 hCoV-19/Denmark/DCGC-17912/2020|EPI_ISL_749640|202 ···················· ···················· ···················· ···················· ···················· ····················
AA 826 hCoV-19/Denmark/DCGC-17913/2020|EPI_ISL_749641|202 ···················· ···················· ···················· ···················· ···················· ····················
AA 827 hCoV-19/Denmark/DCGC-17916/2020|EPI_ISL_749642|202 ···················· ·A·················· ···················· ···················· ···················· ····················
AA 828 hCoV-19/Denmark/DCGC-17918/2020|EPI_ISL_749643|202 ···················· ···················· ···················· ···················· ···················· ····················
AA 829 hCoV-19/Denmark/DCGC-17919/2020|EPI_ISL_749644|202 ···················· ···················· ···················· ···················· ···················· ····················
AA 830 hCoV-19/Denmark/DCGC-17922/2020|EPI_ISL_749645|202 ···················· ···················· ···················· ···················· ···················· ····················
AA 831 hCoV-19/Denmark/DCGC-17923/2020|EPI_ISL_749646|202 ···················· ···················· ···················· ···················· ···················· ····················
AA 832 hCoV-19/Denmark/DCGC-17925/2020|EPI_ISL_749647|202 ···················· ···················· ···················· ···················· ···················· ····················
AA 833 hCoV-19/Denmark/DCGC-17926/2020|EPI_ISL_749648|202 ···················· ···················· ···················· ···················· ···················· ····················
AA 834 hCoV-19/Denmark/DCGC-17927/2020|EPI_ISL_749649|202 ···················· ···················· ···················· ···················· ···················· ····················
AA 835 hCoV-19/Denmark/DCGC-17930/2020|EPI_ISL_749650|202 ···················· ···················· ···················· ···················· ···················· ····················
AA 836 hCoV-19/Denmark/DCGC-17931/2020|EPI_ISL_749651|202 ···················· ···················· ···················· ···················· ···················· ····················
AA 837 hCoV-19/Denmark/DCGC-17933/2020|EPI_ISL_749652|202 ···················· ···················· ···················· ···················· ···················· ····················
AA 838 hCoV-19/Denmark/DCGC-17934/2020|EPI_ISL_749653|202 ···················· ···················· ···················· ···················· ···················· ····················
AA 839 hCoV-19/Denmark/DCGC-17935/2020|EPI_ISL_749654|202 ···················· ···················· ···················· ···················· ···················· ····················
AA 840 hCoV-19/Denmark/DCGC-17937/2020|EPI_ISL_749655|202 ···················· ···················· ···················· ···················· ···················· ····················
AA 841 hCoV-19/Denmark/DCGC-17938/2020|EPI_ISL_749656|202 ···················· ···················· ···················· ···················· ···················· ····················
AA 842 hCoV-19/Denmark/DCGC-17940/2020|EPI_ISL_749657|202 ···················· ···················· ···················· ···················· ···················· ····················
AA 843 hCoV-19/Denmark/DCGC-17942/2020|EPI_ISL_749658|202 ···················· ···················· ···················· ···················· ···················· ····················
AA 844 hCoV-19/Denmark/DCGC-17943/2020|EPI_ISL_749659|202 ···················· ···················· ···················· ···················· ···················· ····················
AA 845 hCoV-19/Denmark/DCGC-17944/2020|EPI_ISL_749660|202 ···················· ···················· ···················· ···················· ···················· ····················
AA 846 hCoV-19/Denmark/DCGC-17948/2020|EPI_ISL_749661|202 ···················· ···················· ···················· ···················· ···················· ····················
AA 847 hCoV-19/Denmark/DCGC-17951/2020|EPI_ISL_749662|202 ···················· ···················· ···················· ···················· ···················· ····················
AA 848 hCoV-19/Denmark/DCGC-17953/2020|EPI_ISL_749663|202 ···················· ···················· ···················· ···················· ···················· ····················
AA 849 hCoV-19/Denmark/DCGC-17954/2020|EPI_ISL_749664|202 ···················· ···················· ···················· ···················· ···················· ····················
AA 850 hCoV-19/Denmark/DCGC-17955/2020|EPI_ISL_749665|202 ···················· ·A·················· ···················· ···················· ···················· ··A·················
AA 851 hCoV-19/Denmark/DCGC-17958/2020|EPI_ISL_749666|202 ···················· ···················· ···················· ···················· ···················· ····················
AA 852 hCoV-19/Denmark/DCGC-17961/2020|EPI_ISL_749667|202 ···················· ···················· ···················· ···················· ···················· ····················
AA 853 hCoV-19/Denmark/DCGC-17962/2020|EPI_ISL_749668|202 ···················· ···················· ···················· ···················· ···················· ····················
AA 854 hCoV-19/Denmark/DCGC-17964/2020|EPI_ISL_749669|202 ···················· ···················· ···················· ···················· ···················· ····················
AA 855 hCoV-19/Denmark/DCGC-17966/2020|EPI_ISL_749670|202 ···················· ···················· ···················· ···················· ···················· ····················
AA 856 hCoV-19/Denmark/DCGC-17971/2020|EPI_ISL_749671|202 ···················· ···················· ···················· ···················· ···················· ····················
AA 857 hCoV-19/Denmark/DCGC-17972/2020|EPI_ISL_749672|202 ···················· ···················· ···················· ···················· ···················· ····················
AA 858 hCoV-19/Denmark/DCGC-17983/2020|EPI_ISL_749680|202 ···················· ···················· ···················· ···················· ···················· ····················
AA 859 hCoV-19/Denmark/DCGC-17984/2020|EPI_ISL_749681|202 ···················· ···················· ···················· ···················· ···················· ····················
AA 860 hCoV-19/Denmark/DCGC-17985/2020|EPI_ISL_749682|202 ···················· ···················· ···················· ···················· ···················· ····················
AA 861 hCoV-19/Denmark/DCGC-17988/2020|EPI_ISL_749683|202 ···················· ···················· ···················· ···················· ···················· ····················
AA 862 hCoV-19/Denmark/DCGC-17989/2020|EPI_ISL_749684|202 ···················· ···················· ···················· ···················· ···················· ····················
AA 863 hCoV-19/Denmark/DCGC-17991/2020|EPI_ISL_749685|202 ···················· ···················· ···················· ···················· ···················· ····················
AA 864 hCoV-19/Denmark/DCGC-17992/2020|EPI_ISL_749686|202 ···················· ···················· ···················· ···················· ···················· ····················
AA 865 hCoV-19/Denmark/DCGC-17994/2020|EPI_ISL_749687|202 ···················· ···················· ···················· ···················· ···················· ····················
AA 866 hCoV-19/Denmark/DCGC-17995/2020|EPI_ISL_749688|202 ···················· ···················· ···················· ···················· ···················· ····················
AA 867 hCoV-19/Denmark/DCGC-17997/2020|EPI_ISL_749689|202 ···················· ···················· ···················· ···················· ···················· ····················
AA 868 hCoV-19/Denmark/DCGC-17998/2020|EPI_ISL_749690|202 ···················· ···················· ···················· ···················· ···················· ····················
AA 869 hCoV-19/Denmark/DCGC-17999/2020|EPI_ISL_749691|202 ···················· ···················· ···················· ···················· ···················· ····················
AA 870 hCoV-19/Denmark/DCGC-18001/2020|EPI_ISL_749692|202 ···················· ···················· ···················· ···················· ···················· ····················
AA 871 hCoV-19/Denmark/DCGC-18003/2020|EPI_ISL_749693|202 ···················· ···················· ···················· ···················· ···················· ····················
AA 872 hCoV-19/Denmark/DCGC-18004/2020|EPI_ISL_749694|202 ···················· ···················· ···················· ···················· ···················· ····················
AA 873 hCoV-19/Denmark/DCGC-18005/2020|EPI_ISL_749695|202 ···················· ···················· ···················· ···················· ···················· ····················
AA 874 hCoV-19/Denmark/DCGC-18006/2020|EPI_ISL_749696|202 ···················· ···················· ···················· ···················· ···················· ····················
AA 875 hCoV-19/Denmark/DCGC-18007/2020|EPI_ISL_749697|202 ···················· ···················· ···················· ···················· ···················· ····················
AA 876 hCoV-19/Denmark/DCGC-18009/2020|EPI_ISL_749698|202 ···················· ···················· ···················· ···················· ···················· ····················
AA 877 hCoV-19/Denmark/DCGC-18010/2020|EPI_ISL_749699|202 ···················· ···················· ···················· ···················· ···················· ····················
AA 878 hCoV-19/Denmark/DCGC-16882/2020|EPI_ISL_750620|202 ···················· ···················· ···················· ···················· ···················· ····················
AA 879 hCoV-19/Denmark/DCGC-16883/2020|EPI_ISL_750621|202 ···················· ···················· ···················· ···················· ···················· ····················

| | | | | | | | |
|---|---|---|---|---|---|---|---|
| AA | 880 hCoV-19/Denmark/DCGC-16884/2020\|EPI_ISL_750622\|202 | .................... | .................... | ..................... | .................... | .................... | .................... |
| AA | 881 hCoV-19/Denmark/DCGC-16885/2020\|EPI_ISL_750623\|202 | .................... | .................... | ..................... | .................... | .................... | .................... |
| AA | 882 hCoV-19/Denmark/DCGC-16889/2020\|EPI_ISL_750624\|202 | .................... | .................... | ..................... | .................... | .................... | .................... |
| AA | 883 hCoV-19/Denmark/DCGC-16892/2020\|EPI_ISL_750625\|202 | .................... | .................... | ..................... | .................... | .................... | .................... |
| AA | 884 hCoV-19/Denmark/DCGC-16893/2020\|EPI_ISL_750626\|202 | .................... | .................... | ..................... | .................... | .................... | .................... |
| AA | 885 hCoV-19/Denmark/DCGC-16894/2020\|EPI_ISL_750627\|202 | .................... | .................... | ..................... | .................... | .................... | .................... |
| AA | 886 hCoV-19/Denmark/DCGC-16895/2020\|EPI_ISL_750628\|202 | .................... | .A.................. | ..................... | .................... | .................... | ..A................. |
| AA | 887 hCoV-19/Denmark/DCGC-16896/2020\|EPI_ISL_750629\|202 | .................... | .................... | ..................... | .................... | .................... | .................... |
| AA | 888 hCoV-19/Denmark/DCGC-16897/2020\|EPI_ISL_750630\|202 | .................... | .................... | ..................... | .................... | .................... | .................... |
| AA | 889 hCoV-19/Denmark/DCGC-16898/2020\|EPI_ISL_750631\|202 | .................... | .................... | ..................... | .................... | .................... | .................... |
| AA | 890 hCoV-19/Denmark/DCGC-16900/2020\|EPI_ISL_750632\|202 | .................... | .................... | ..................... | .................... | .................... | .................... |
| AA | 891 hCoV-19/Denmark/DCGC-16901/2020\|EPI_ISL_750633\|202 | .................... | .................... | ..................... | .................... | .................... | .................... |
| AA | 892 hCoV-19/Denmark/DCGC-16902/2020\|EPI_ISL_750634\|202 | .................... | .................... | ..................... | .................... | .................... | .................... |
| AA | 893 hCoV-19/Denmark/DCGC-16904/2020\|EPI_ISL_750635\|202 | .................... | .................... | ..................... | .................... | .................... | .................... |
| AA | 894 hCoV-19/Denmark/DCGC-16907/2020\|EPI_ISL_750636\|202 | .................... | .................... | ..................... | .................... | .................... | .................... |
| AA | 895 hCoV-19/Denmark/DCGC-16909/2020\|EPI_ISL_750637\|202 | .................... | .................... | ..................... | .................... | .................... | .................... |
| AA | 896 hCoV-19/Denmark/DCGC-16910/2020\|EPI_ISL_750638\|202 | .................... | .................... | ..................... | .................... | .................... | .................... |
| AA | 897 hCoV-19/Denmark/DCGC-16912/2020\|EPI_ISL_750639\|202 | .................... | .................... | ..................... | .................... | .................... | .................... |
| AA | 898 hCoV-19/Denmark/DCGC-16915/2020\|EPI_ISL_750640\|202 | .................... | .................... | ..................... | .................... | .................... | .................... |
| AA | 899 hCoV-19/Denmark/DCGC-16917/2020\|EPI_ISL_750641\|202 | .................... | .................... | ..................... | .................... | .................... | .................... |
| AA | 900 hCoV-19/Denmark/DCGC-16921/2020\|EPI_ISL_750642\|202 | .................... | .................... | ..................... | .................... | .................... | .................... |
| AA | 901 hCoV-19/Denmark/DCGC-16922/2020\|EPI_ISL_750643\|202 | .................... | .................... | ..................... | .................... | .................... | .................... |
| AA | 902 hCoV-19/Denmark/DCGC-16923/2020\|EPI_ISL_750644\|202 | .................... | .................... | ..................... | .................... | .................... | .................... |
| AA | 903 hCoV-19/Denmark/DCGC-16924/2020\|EPI_ISL_750645\|202 | .................... | .................... | ..................... | .................... | .................... | .................... |
| AA | 904 hCoV-19/Denmark/DCGC-16925/2020\|EPI_ISL_750646\|202 | .................... | .................... | ..................... | .................... | .................... | .................... |
| AA | 905 hCoV-19/Denmark/DCGC-16927/2020\|EPI_ISL_750647\|202 | .................... | .................... | ..................... | .................... | .................... | .................... |
| AA | 906 hCoV-19/Denmark/DCGC-16928/2020\|EPI_ISL_750648\|202 | .................... | .................... | ..................... | .................... | .................... | .................... |
| AA | 907 hCoV-19/Denmark/DCGC-16929/2020\|EPI_ISL_750649\|202 | .................... | .................... | ..................... | .................... | .................... | .................... |
| AA | 908 hCoV-19/Denmark/DCGC-16932/2020\|EPI_ISL_750650\|202 | .................... | .................... | ..................... | .................... | .................... | .................... |
| AA | 909 hCoV-19/Denmark/DCGC-16933/2020\|EPI_ISL_750651\|202 | .................... | .................... | ..................... | .................... | .................... | .................... |
| AA | 910 hCoV-19/Denmark/DCGC-16935/2020\|EPI_ISL_750652\|202 | .................... | .................... | ..................... | .................... | .................... | .................... |
| AA | 911 hCoV-19/Denmark/DCGC-16936/2020\|EPI_ISL_750653\|202 | .................... | .................... | ..................... | .................... | .................... | .................... |
| AA | 912 hCoV-19/Denmark/DCGC-16938/2020\|EPI_ISL_750654\|202 | .................... | .................... | ..................... | .................... | .................... | .................... |
| AA | 913 hCoV-19/Denmark/DCGC-16939/2020\|EPI_ISL_750655\|202 | .................... | .................... | ..................... | .................... | .................... | .................... |
| AA | 914 hCoV-19/Denmark/DCGC-16941/2020\|EPI_ISL_750656\|202 | .................... | .................... | ..................... | .................... | .................... | .................... |
| AA | 915 hCoV-19/Denmark/DCGC-16944/2020\|EPI_ISL_750657\|202 | .................... | .................... | ..................... | .................... | .................... | .................... |
| AA | 916 hCoV-19/Denmark/DCGC-16945/2020\|EPI_ISL_750658\|202 | .................... | .................... | ..................... | .................... | .................... | .................... |
| AA | 917 hCoV-19/Denmark/DCGC-16946/2020\|EPI_ISL_750659\|202 | .................... | .................... | ..................... | .................... | .................... | .................... |
| AA | 918 hCoV-19/Denmark/DCGC-16947/2020\|EPI_ISL_750660\|202 | .................... | .................... | ..................... | .................... | .................... | .................... |
| AA | 919 hCoV-19/Denmark/DCGC-16948/2020\|EPI_ISL_750661\|202 | .................... | .A.................. | ..................... | .................... | .................... | .................... |
| AA | 920 hCoV-19/Denmark/DCGC-16949/2020\|EPI_ISL_750662\|202 | .................... | .................... | ..................... | .................... | .................... | .................... |
| AA | 921 hCoV-19/Denmark/DCGC-16952/2020\|EPI_ISL_750663\|202 | .................... | .................... | ..................... | .................... | .................... | .................... |
| AA | 922 hCoV-19/Denmark/DCGC-16963/2020\|EPI_ISL_750670\|202 | .................... | .................... | ..................... | .................... | .................... | .................... |
| AA | 923 hCoV-19/Denmark/DCGC-16964/2020\|EPI_ISL_750671\|202 | .................... | .................... | ..................... | .................... | .................... | .................... |
| AA | 924 hCoV-19/Denmark/DCGC-16965/2020\|EPI_ISL_750672\|202 | .................... | .................... | ..................... | .................... | .................... | .................... |
| AA | 925 hCoV-19/Denmark/DCGC-16966/2020\|EPI_ISL_750673\|202 | .................... | .................... | ..................... | .................... | .................... | .................... |
| AA | 926 hCoV-19/Denmark/DCGC-16967/2020\|EPI_ISL_750674\|202 | .................... | .................... | ..................... | .................... | .................... | .................... |
| AA | 927 hCoV-19/Denmark/DCGC-16970/2020\|EPI_ISL_750675\|202 | .................... | .................... | ..................... | .................... | .................... | .................... |
| AA | 928 hCoV-19/Denmark/DCGC-16971/2020\|EPI_ISL_750676\|202 | .................... | .................... | ..................... | .................... | .................... | .................... |
| AA | 929 hCoV-19/Denmark/DCGC-16972/2020\|EPI_ISL_750677\|202 | .................... | .................... | ..................... | .................... | .................... | .................... |
| AA | 930 hCoV-19/Denmark/DCGC-16973/2020\|EPI_ISL_750678\|202 | .................... | .................... | ..................... | .................... | .................... | .................... |
| AA | 931 hCoV-19/Denmark/DCGC-16974/2020\|EPI_ISL_750679\|202 | .................... | .................... | ..................... | .................... | .................... | .................... |
| AA | 932 hCoV-19/Denmark/DCGC-16975/2020\|EPI_ISL_750680\|202 | .................... | .................... | ..................... | .................... | .................... | .................... |
| AA | 933 hCoV-19/Denmark/DCGC-16977/2020\|EPI_ISL_750681\|202 | .................... | .................... | ..................... | .................... | .................... | .................... |
| AA | 934 hCoV-19/Denmark/DCGC-16978/2020\|EPI_ISL_750682\|202 | .................... | .................... | ..................... | .................... | .................... | .................... |
| AA | 935 hCoV-19/Denmark/DCGC-16979/2020\|EPI_ISL_750683\|202 | .................... | .................... | ..................... | .................... | .................... | .................... |
| AA | 936 hCoV-19/Denmark/DCGC-16980/2020\|EPI_ISL_750684\|202 | .................... | .................... | ..................... | .................... | .................... | .................... |
| AA | 937 hCoV-19/Denmark/DCGC-16981/2020\|EPI_ISL_750685\|202 | .................... | .................... | ..................... | .................... | .................... | .................... |
| AA | 938 hCoV-19/Denmark/DCGC-16982/2020\|EPI_ISL_750686\|202 | .................... | .................... | ..................... | .................... | .................... | .................... |
| AA | 939 hCoV-19/Denmark/DCGC-16983/2020\|EPI_ISL_750687\|202 | .................... | .................... | ..................... | .................... | .................... | .................... |
| AA | 940 hCoV-19/Denmark/DCGC-16984/2020\|EPI_ISL_750688\|202 | .................... | .................... | ..................... | .................... | .................... | .................... |
| AA | 941 hCoV-19/Denmark/DCGC-16985/2020\|EPI_ISL_750689\|202 | .................... | .................... | ..................... | .................... | .................... | .................... |
| AA | 942 hCoV-19/Denmark/DCGC-16986/2020\|EPI_ISL_750690\|202 | .................... | .................... | ..................... | .................... | .................... | .................... |
| AA | 943 hCoV-19/Denmark/DCGC-16987/2020\|EPI_ISL_750691\|202 | .................... | .................... | ..................... | .................... | .................... | .................... |
| AA | 944 hCoV-19/Denmark/DCGC-16988/2020\|EPI_ISL_750692\|202 | .................... | .................... | ..................... | .................... | .................... | .................... |
| AA | 945 hCoV-19/Denmark/DCGC-16989/2020\|EPI_ISL_750693\|202 | .................... | .................... | ..................... | .................... | .................... | .................... |
| AA | 946 hCoV-19/Denmark/DCGC-16990/2020\|EPI_ISL_750694\|202 | .................... | .................... | ..................... | .................... | .................... | .................... |
| AA | 947 hCoV-19/Denmark/DCGC-16991/2020\|EPI_ISL_750695\|202 | .................... | .................... | ..................... | .................... | .................... | .................... |
| AA | 948 hCoV-19/Denmark/DCGC-16992/2020\|EPI_ISL_750696\|202 | .................... | .................... | ..................... | .................... | .................... | .................... |
| AA | 949 hCoV-19/Denmark/DCGC-16993/2020\|EPI_ISL_750697\|202 | .................... | .................... | ..................... | .................... | .................... | .................... |
| AA | 950 hCoV-19/Denmark/DCGC-16997/2020\|EPI_ISL_750698\|202 | .................... | .................... | ..................... | .................... | .................... | .................... |
| AA | 951 hCoV-19/Denmark/DCGC-16998/2020\|EPI_ISL_750699\|202 | .................... | .................... | ..................... | .................... | .................... | .................... |
| AA | 952 hCoV-19/USA/ME-HETL-J0385/2020\|EPI_ISL_755500\|2020 | .................... | .................... | ..................... | .................... | .................... | .................... |
| AA | 953 hCoV-19/USA/ME-HETL-J0388/2020\|EPI_ISL_755502\|2020 | .................... | .................... | ..................... | .................... | .................... | .................... |
| AA | 954 hCoV-19/USA/ME-HETL-J0389/2020\|EPI_ISL_755503\|2020 | .................... | .A.................. | ..................... | .................... | .................... | .................... |
| AA | 955 hCoV-19/USA/ME-HETL-J0395/2020\|EPI_ISL_755507\|2020 | .................... | .................... | ..................... | .................... | .................... | .................... |
| AA | 956 hCoV-19/USA/ME-HETL-J0397/2020\|EPI_ISL_755508\|2020 | .................... | .................... | ..................... | .................... | .................... | .................... |
| AA | 957 hCoV-19/USA/ME-HETL-J0399/2020\|EPI_ISL_755509\|2020 | .................... | .................... | ..................... | .................... | .................... | .................... |
| AA | 958 hCoV-19/USA/ME-HETL-J0400/2020\|EPI_ISL_755510\|2020 | .................... | .................... | ..................... | .................... | .................... | .................... |
| AA | 959 hCoV-19/USA/ME-HETL-J0402/2020\|EPI_ISL_755511\|2020 | .................... | .................... | ..................... | .................... | .................... | .................... |

```
AA   960 hCoV-19/USA/ME-HETL-J0403/2020|EPI_ISL_755512|2020   ····················  ····················  ·····················  ····················  ····················  ····················
AA   961 hCoV-19/USA/ME-HETL-J0404/2020|EPI_ISL_755513|2020   ····················  ····················  ·····················  ····················  ····················  ····················
AA   962 hCoV-19/USA/ME-HETL-J0406/2020|EPI_ISL_755514|2020   ····················  ····················  ·····················  ····················  ····················  ····················
AA   963 hCoV-19/England/MILK-C7AE92/2020|EPI_ISL_756816|20   ····················  ····················  ·····················  ····················  ····················  ····················
AA   964 hCoV-19/England/MILK-C7B909/2020|EPI_ISL_756817|20   ····················  ····················  ·····················  ····················  ····················  ····················
AA   965 hCoV-19/England/MILK-C78A18/2020|EPI_ISL_756818|20   ····················  ····················  ·····················  ····················  ····················  ····················
AA   966 hCoV-19/England/MILK-C7B453/2020|EPI_ISL_756822|20   ····················  ····················  ·····················  ····················  ····················  ····················
AA   967 hCoV-19/England/QEUH-C7C7FF/2020|EPI_ISL_756824|20   ····················  ····················  ·····················  ····················  ····················  ····················
AA   968 hCoV-19/England/MILK-C7B3A1/2020|EPI_ISL_756825|20   ···T················  ····················  ·····················  ····················  ····················  ····················
AA   969 hCoV-19/England/MILK-C7B392/2020|EPI_ISL_756828|20   ····················  ····················  ·····················  ····················  ····················  ····················
AA   970 hCoV-19/England/QEUH-C8027F/2020|EPI_ISL_756835|20   ····················  ····················  ·····················  ····················  ····················  ····················
AA   971 hCoV-19/England/MILK-C784DE/2020|EPI_ISL_756837|20   ····················  ····················  ·····················  ····················  ····················  ····················
AA   972 hCoV-19/England/MILK-C78A36/2020|EPI_ISL_756841|20   ····················  ····················  ·····················  ····················  ····················  ····················
AA   973 hCoV-19/England/MILK-C7824D/2020|EPI_ISL_756843|20   ····················  ····················  ·····················  ····················  ····················  ····················
AA   974 hCoV-19/England/MILK-C787B7/2020|EPI_ISL_756844|20   ····················  ····················  ·····················  ····················  ····················  ····················
AA   975 hCoV-19/Denmark/DCGC-20678/2020|EPI_ISL_759000|202   ····················  ··G·················  ·····················  ····················  ····················  ····················
AA   976 hCoV-19/Denmark/DCGC-20712/2020|EPI_ISL_759001|202   ····················  ····················  ·····················  ····················  ····················  ····················
AA   977 hCoV-19/Denmark/DCGC-20722/2020|EPI_ISL_759002|202   ····················  ····················  ·····················  ····················  ····················  ····················
AA   978 hCoV-19/Denmark/DCGC-20737/2020|EPI_ISL_759003|202   ····················  ····················  ·····················  ····················  ····················  ····················
AA   979 hCoV-19/Denmark/DCGC-20788/2020|EPI_ISL_759004|202   ····················  ····················  ·····················  ····················  ····················  ····················
AA   980 hCoV-19/Denmark/DCGC-20794/2020|EPI_ISL_759005|202   ····················  ··G·················  ·····················  ····················  ····················  ····················
AA   981 hCoV-19/Denmark/DCGC-20798/2020|EPI_ISL_759006|202   ····················  ····················  ·····················  ····················  ····················  ····················
AA   982 hCoV-19/Denmark/DCGC-20811/2020|EPI_ISL_759007|202   ····················  ····················  ·····················  ····················  ···C················  ····················
AA   983 hCoV-19/Denmark/DCGC-20815/2020|EPI_ISL_759008|202   ····················  ····················  ·····················  ····················  ···C················  ····················
AA   984 hCoV-19/Denmark/DCGC-20831/2020|EPI_ISL_759009|202   ····················  ····················  ·····················  ····················  ···C················  ····················
AA   985 hCoV-19/Denmark/DCGC-20892/2020|EPI_ISL_759010|202   ····················  ····················  ·····················  ····················  ····················  ····················
AA   986 hCoV-19/Denmark/DCGC-20901/2020|EPI_ISL_759011|202   ····················  ····················  ·····················  ····················  ··· T···············  ····················
AA   987 hCoV-19/Denmark/DCGC-20903/2020|EPI_ISL_759012|202   ····················  ····················  ·····················  ····················  ····················  ····················
AA   988 hCoV-19/Denmark/DCGC-20908/2020|EPI_ISL_759013|202   ····················  ····················  ·····················  ····················  ····················  ····················
AA   989 hCoV-19/Denmark/DCGC-20915/2020|EPI_ISL_759014|202   ····················  ····················  ·····················  ····················  ····················  ····················
AA   990 hCoV-19/Denmark/DCGC-20929/2020|EPI_ISL_759015|202   ····················  ····················  ·····················  ····················  ···C················  ····················
AA   991 hCoV-19/Denmark/DCGC-20962/2020|EPI_ISL_759016|202   ····················  ····················  ·····················  ····················  ···C················  ····················
AA   992 hCoV-19/Denmark/DCGC-20963/2020|EPI_ISL_759017|202   ····················  ····················  ·····················  ····················  ····················  ····················
AA   993 hCoV-19/Denmark/DCGC-20981/2020|EPI_ISL_759018|202   ····················  ····················  ·····················  ····················  ····················  ····················
AA   994 hCoV-19/Denmark/DCGC-20990/2020|EPI_ISL_759019|202   ····················  ····················  ·····················  ····················  ····················  ····················
AA   995 hCoV-19/Denmark/DCGC-20994/2020|EPI_ISL_759020|202   ····················  ····················  ·····················  ····················  ····················  ····················
AA   996 hCoV-19/Denmark/DCGC-21021/2020|EPI_ISL_759021|202   ····················  ····················  ·····················  ····················  ····················  ····················
AA   997 hCoV-19/Denmark/DCGC-21045/2020|EPI_ISL_759022|202   ····················  ····················  ·····················  ····················  ····················  ····················
AA   998 hCoV-19/Denmark/DCGC-21052/2020|EPI_ISL_759023|202   ····················  ····················  ·····················  ····················  ····················  ····················
AA   999 hCoV-19/Denmark/DCGC-21067/2020|EPI_ISL_759024|202   ····················  ····················  ·····················  ····················  ····················  ····················
AA  1000 hCoV-19/Denmark/DCGC-21080/2020|EPI_ISL_759025|202   ····················  ····················  ·····················  ····················  ····················  ····················
AA  1001 hCoV-19/Denmark/DCGC-21081/2020|EPI_ISL_759026|202   ····················  ····················  ·····················  ····················  ····················  ····················
AA  1002 hCoV-19/Denmark/DCGC-21107/2020|EPI_ISL_759027|202   ····················  ····················  ·····················  ····················  ····················  ····················
AA  1003 hCoV-19/Denmark/DCGC-21165/2020|EPI_ISL_759028|202   ····················  ····················  ·····················  ····················  ····················  ····················
AA  1004 hCoV-19/Denmark/DCGC-21166/2020|EPI_ISL_759029|202   ····················  ····················  ·····················  ····················  ···C················  ····················
AA  1005 hCoV-19/Denmark/DCGC-21168/2020|EPI_ISL_759030|202   ····················  ····················  ·····················  ····················  ····················  ····················
AA  1006 hCoV-19/Denmark/DCGC-21189/2020|EPI_ISL_759031|202   ····················  ····················  ·····················  ····················  ····················  ····················
AA  1007 hCoV-19/Denmark/DCGC-21206/2020|EPI_ISL_759032|202   ····················  ····················  ·····················  ····················  ····················  ····················
AA  1008 hCoV-19/Denmark/DCGC-21213/2020|EPI_ISL_759033|202   ····················  ····················  ·····················  ····················  ····················  ····················
AA  1009 hCoV-19/Denmark/DCGC-21235/2020|EPI_ISL_759034|202   ····················  ····················  ·····················  ····················  ····················  ····················
AA  1010 hCoV-19/Denmark/DCGC-21240/2020|EPI_ISL_759035|202   ····················  ····················  ·····················  ····················  ····················  ····················
AA  1011 hCoV-19/Denmark/DCGC-21263/2020|EPI_ISL_759036|202   ····················  ····················  ·····················  ····················  ····················  ····················
AA  1012 hCoV-19/Denmark/DCGC-21280/2020|EPI_ISL_759037|202   ····················  ····················  ·····················  ····················  ····················  ····················
AA  1013 hCoV-19/Denmark/DCGC-21305/2020|EPI_ISL_759038|202   ····················  ····················  ·····················  ····················  ····················  ····················
AA  1014 hCoV-19/Denmark/DCGC-21311/2020|EPI_ISL_759039|202   ····················  ····················  ·····················  ····················  ····················  ····················
AA  1015 hCoV-19/Denmark/DCGC-21336/2020|EPI_ISL_759040|202   ····················  ····················  ·····················  ····················  ····················  ····················
AA  1016 hCoV-19/Denmark/DCGC-20493/2020|EPI_ISL_759046|202   ····················  ····················  ·····················  ····················  ····················  ····················
AA  1017 hCoV-19/Denmark/DCGC-20544/2020|EPI_ISL_759047|202   ····················  ·A··················  ·····················  ····················  ····················  ··A·················
AA  1018 hCoV-19/Denmark/DCGC-20684/2020|EPI_ISL_759048|202   ····················  ·A··················  ·····················  ····················  ····················  ··A·················
AA  1019 hCoV-19/Denmark/DCGC-20779/2020|EPI_ISL_759049|202   ····················  ····················  ·····················  ····················  ····················  ····················
AA  1020 hCoV-19/Denmark/DCGC-20985/2020|EPI_ISL_759050|202   ····················  ·A··················  ·····················  ····················  ····················  ··A·················
AA  1021 hCoV-19/Denmark/DCGC-21014/2020|EPI_ISL_759051|202   ····················  ·A··················  ·····················  ····················  ····················  ··A·················
AA  1022 hCoV-19/Denmark/DCGC-21043/2020|EPI_ISL_759052|202   ····················  ····················  ·····················  ····················  ····················  ····················
AA  1023 hCoV-19/Denmark/DCGC-21089/2020|EPI_ISL_759054|202   ····················  ·A··················  ·····················  ····················  ····················  ··A·················
AA  1024 hCoV-19/Denmark/DCGC-21171/2020|EPI_ISL_759055|202   ····················  ····················  ·····················  ····················  ····················  ····················
AA  1025 hCoV-19/Denmark/DCGC-18933/2020|EPI_ISL_748400|202   ····················  ····················  ·····················  ····················  ····················  ····················
AA  1026 hCoV-19/Denmark/DCGC-18936/2020|EPI_ISL_748401|202   ····················  ····················  ·····················  ····················  ····················  ····················
AA  1027 hCoV-19/Denmark/DCGC-18942/2020|EPI_ISL_748402|202   ····················  ····················  ·····················  ····················  ····················  ····················
AA  1028 hCoV-19/Denmark/DCGC-18953/2020|EPI_ISL_748403|202   ····················  ····················  ·····················  ····················  ····················  ····················
AA  1029 hCoV-19/Denmark/DCGC-18968/2020|EPI_ISL_748404|202   ····················  ····················  ·····················  ····················  ····················  ····················
AA  1030 hCoV-19/Denmark/DCGC-18977/2020|EPI_ISL_748405|202   ····················  ····················  ·····················  ····················  ····················  ····················
AA  1031 hCoV-19/Denmark/DCGC-18996/2020|EPI_ISL_748406|202   ····················  ····················  ·····················  ····················  ····················  ····················
AA  1032 hCoV-19/Denmark/DCGC-19007/2020|EPI_ISL_748407|202   ····················  ····················  ·····················  ····················  ····················  ····················
AA  1033 hCoV-19/Denmark/DCGC-19029/2020|EPI_ISL_748408|202   ····················  ····················  ·····················  ····················  ····················  ····················
AA  1034 hCoV-19/Denmark/DCGC-19033/2020|EPI_ISL_748409|202   ····················  ····················  ·····················  ····················  ····················  ····················
AA  1035 hCoV-19/Denmark/DCGC-19042/2020|EPI_ISL_748410|202   ····················  ····················  ·····················  ····················  ····················  ····················
AA  1036 hCoV-19/Denmark/DCGC-19046/2020|EPI_ISL_748411|202   ····················  ····················  ·····················  ····················  ····················  ····················
AA  1037 hCoV-19/Denmark/DCGC-19049/2020|EPI_ISL_748412|202   ····················  ····················  ·····················  ····················  ····················  ····················
AA  1038 hCoV-19/Denmark/DCGC-19056/2020|EPI_ISL_748413|202   ····················  ····················  ·····················  ····················  ····················  ····················
AA  1039 hCoV-19/Denmark/DCGC-19057/2020|EPI_ISL_748414|202   ····················  ····················  ·····················  ····················  ····················  ····················
```

| | | | | | | | |
|---|---|---|---|---|---|---|---|
| AA | 1040 hCoV-19/Denmark/DCGC-19099/2020\|EPI_ISL_748415\|202 | ···················· | ···················· | ···················· | ···················· | ···················· | ···················· |
| AA | 1041 hCoV-19/Denmark/DCGC-19100/2020\|EPI_ISL_748416\|202 | ···················· | ···················· | ···················· | ···················· | ···················· | ···················· |
| AA | 1042 hCoV-19/Denmark/DCGC-19101/2020\|EPI_ISL_748417\|202 | ···················· | ···················· | ···················· | ···················· | ···················· | ···················· |
| AA | 1043 hCoV-19/Denmark/DCGC-19102/2020\|EPI_ISL_748418\|202 | ···················· | ···················· | ···················· | ···················· | ···················· | ···················· |
| AA | 1044 hCoV-19/Denmark/DCGC-19103/2020\|EPI_ISL_748419\|202 | ···················· | ···················· | ···················· | ···················· | ···················· | ···················· |
| AA | 1045 hCoV-19/Denmark/DCGC-19125/2020\|EPI_ISL_748420\|202 | ···················· | ···················· | ···················· | ···················· | ···················· | ···················· |
| AA | 1046 hCoV-19/Denmark/DCGC-19134/2020\|EPI_ISL_748421\|202 | ···················· | ···················· | ···················· | ···················· | ···················· | ···················· |
| AA | 1047 hCoV-19/Denmark/DCGC-19180/2020\|EPI_ISL_748422\|202 | ···················· | ···················· | ···················· | ···················· | ···················· | ···················· |
| AA | 1048 hCoV-19/Denmark/DCGC-19213/2020\|EPI_ISL_748423\|202 | ···················· | ···················· | ···················· | ···················· | ···················· | ···················· |
| AA | 1049 hCoV-19/Denmark/DCGC-19214/2020\|EPI_ISL_748424\|202 | ···················· | ···················· | ···················· | ···················· | ···················· | ···················· |
| AA | 1050 hCoV-19/Denmark/DCGC-19231/2020\|EPI_ISL_748425\|202 | ···················· | ···················· | ···················· | ···················· | ···················· | ···················· |
| AA | 1051 hCoV-19/Denmark/DCGC-19253/2020\|EPI_ISL_748426\|202 | ···················· | ···················· | ···················· | ···················· | ···················· | ···················· |
| AA | 1052 hCoV-19/Denmark/DCGC-19258/2020\|EPI_ISL_748427\|202 | ···················· | ···················· | ···················· | ···················· | ···················· | ···················· |
| AA | 1053 hCoV-19/Denmark/DCGC-19266/2020\|EPI_ISL_748428\|202 | ···················· | ···················· | ···················· | ···················· | ···················· | ···················· |
| AA | 1054 hCoV-19/Denmark/DCGC-19269/2020\|EPI_ISL_748429\|202 | ···················· | ···················· | ···················· | ···················· | ···················· | ···················· |
| AA | 1055 hCoV-19/Denmark/DCGC-18011/2020\|EPI_ISL_749700\|202 | ···················· | ···················· | ···················· | ···················· | ···················· | ···················· |
| AA | 1056 hCoV-19/Denmark/DCGC-18012/2020\|EPI_ISL_749701\|202 | ···················· | ···················· | ···················· | ···················· | ···················· | ···················· |
| AA | 1057 hCoV-19/Denmark/DCGC-18015/2020\|EPI_ISL_749702\|202 | ···················· | ···················· | ···················· | ···················· | ···················· | ···················· |
| AA | 1058 hCoV-19/Denmark/DCGC-18017/2020\|EPI_ISL_749703\|202 | ···················· | ···················· | ···················· | ···················· | ···················· | ···················· |
| AA | 1059 hCoV-19/Denmark/DCGC-18019/2020\|EPI_ISL_749704\|202 | ···················· | ···················· | ···················· | ···················· | ···················· | ···················· |
| AA | 1060 hCoV-19/Denmark/DCGC-18021/2020\|EPI_ISL_749705\|202 | ···················· | ···················· | ···················· | ···················· | ···················· | ···················· |
| AA | 1061 hCoV-19/Denmark/DCGC-18022/2020\|EPI_ISL_749707\|202 | ···················· | ···················· | ···················· | ···················· | ···················· | ···················· |
| AA | 1062 hCoV-19/Denmark/DCGC-18023/2020\|EPI_ISL_749708\|202 | ···················· | ···················· | ···················· | ···················· | ···················· | ···················· |
| AA | 1063 hCoV-19/Denmark/DCGC-18026/2020\|EPI_ISL_749709\|202 | ···················· | ···················· | ···················· | ···················· | ···················· | ···················· |
| AA | 1064 hCoV-19/Denmark/DCGC-18027/2020\|EPI_ISL_749710\|202 | ···················· | ···················· | ···················· | ···················· | ···················· | ···················· |
| AA | 1065 hCoV-19/Denmark/DCGC-18028/2020\|EPI_ISL_749711\|202 | ···················· | ···················· | ···················· | ···················· | ···················· | ···················· |
| AA | 1066 hCoV-19/Denmark/DCGC-18030/2020\|EPI_ISL_749712\|202 | ···················· | ···················· | ···················· | ···················· | ···················· | ···················· |
| AA | 1067 hCoV-19/Denmark/DCGC-18031/2020\|EPI_ISL_749713\|202 | ···················· | ···················· | ···················· | ···················· | ···················· | ···················· |
| AA | 1068 hCoV-19/Denmark/DCGC-18032/2020\|EPI_ISL_749714\|202 | ···················· | ···················· | ···················· | ···················· | ···················· | ···················· |
| AA | 1069 hCoV-19/Denmark/DCGC-18033/2020\|EPI_ISL_749715\|202 | ···················· | ···················· | ···················· | ···················· | ···················· | ···················· |
| AA | 1070 hCoV-19/Denmark/DCGC-18037/2020\|EPI_ISL_749716\|202 | ···················· | ···················· | ···················· | ···················· | ···················· | ···················· |
| AA | 1071 hCoV-19/Denmark/DCGC-18038/2020\|EPI_ISL_749717\|202 | ···················· | ···················· | ···················· | ···················· | ···················· | ···················· |
| AA | 1072 hCoV-19/Denmark/DCGC-18039/2020\|EPI_ISL_749718\|202 | ···················· | ···················· | ···················· | ···················· | ···················· | ···················· |
| AA | 1073 hCoV-19/Denmark/DCGC-18044/2020\|EPI_ISL_749720\|202 | ···················· | ···················· | ···················· | ···················· | ···················· | ···················· |
| AA | 1074 hCoV-19/Denmark/DCGC-18046/2020\|EPI_ISL_749721\|202 | ···················· | ···················· | ···················· | ···················· | ···················· | ···················· |
| AA | 1075 hCoV-19/Denmark/DCGC-18047/2020\|EPI_ISL_749722\|202 | ···················· | ···················· | ···················· | ···················· | ···················· | ···················· |
| AA | 1076 hCoV-19/Denmark/DCGC-18048/2020\|EPI_ISL_749723\|202 | ···················· | ···················· | ···················· | ···················· | ···················· | ···················· |
| AA | 1077 hCoV-19/Denmark/DCGC-18049/2020\|EPI_ISL_749724\|202 | ···················· | ···················· | ···················· | ···················· | ···················· | ···················· |
| AA | 1078 hCoV-19/Denmark/DCGC-18050/2020\|EPI_ISL_749725\|202 | ···················· | ···················· | ···················· | ···················· | ···················· | ···················· |
| AA | 1079 hCoV-19/Denmark/DCGC-18052/2020\|EPI_ISL_749726\|202 | ···················· | ···················· | ···················· | ···················· | ···················· | ···················· |
| AA | 1080 hCoV-19/Denmark/DCGC-18055/2020\|EPI_ISL_749727\|202 | ···················· | ···················· | ···················· | ···················· | ···················· | ···················· |
| AA | 1081 hCoV-19/Denmark/DCGC-18056/2020\|EPI_ISL_749728\|202 | ···················· | ···················· | ···················· | ···················· | ···················· | ···················· |
| AA | 1082 hCoV-19/Denmark/DCGC-18057/2020\|EPI_ISL_749729\|202 | ···················· | ···················· | ···················· | ···················· | ···················· | ···················· |
| AA | 1083 hCoV-19/Denmark/DCGC-18058/2020\|EPI_ISL_749730\|202 | ···················· | ···················· | ···················· | ···················· | ···················· | ···················· |
| AA | 1084 hCoV-19/Denmark/DCGC-18059/2020\|EPI_ISL_749731\|202 | ···················· | ···················· | ···················· | ···················· | ···················· | ···················· |
| AA | 1085 hCoV-19/Denmark/DCGC-18060/2020\|EPI_ISL_749732\|202 | ···················· | ···················· | ···················· | ···················· | ···················· | ···················· |
| AA | 1086 hCoV-19/Denmark/DCGC-18063/2020\|EPI_ISL_749733\|202 | ···················· | ···················· | ···················· | ···················· | ···················· | ···················· |
| AA | 1087 hCoV-19/Denmark/DCGC-18065/2020\|EPI_ISL_749734\|202 | ···················· | ···················· | ···················· | ···················· | ···················· | ···················· |
| AA | 1088 hCoV-19/Denmark/DCGC-18066/2020\|EPI_ISL_749735\|202 | ···················· | ···················· | ···················· | ···················· | ···················· | ···················· |
| AA | 1089 hCoV-19/Denmark/DCGC-18067/2020\|EPI_ISL_749736\|202 | ···················· | ···················· | ···················· | ···················· | ···················· | ···················· |
| AA | 1090 hCoV-19/Denmark/DCGC-18071/2020\|EPI_ISL_749737\|202 | ···················· | ···················· | ···················· | ···················· | ···················· | ···················· |
| AA | 1091 hCoV-19/Denmark/DCGC-18072/2020\|EPI_ISL_749738\|202 | ···················· | ···················· | ···················· | ···················· | ···················· | ···················· |
| AA | 1092 hCoV-19/Denmark/DCGC-18074/2020\|EPI_ISL_749739\|202 | ···················· | ···················· | ···················· | ···················· | ···················· | ···················· |
| AA | 1093 hCoV-19/Denmark/DCGC-18075/2020\|EPI_ISL_749740\|202 | ···················· | ···················· | ···················· | ···················· | ···················· | ···················· |
| AA | 1094 hCoV-19/Denmark/DCGC-18076/2020\|EPI_ISL_749741\|202 | ···················· | ···················· | ···················· | ···················· | ···················· | ···················· |
| AA | 1095 hCoV-19/Denmark/DCGC-18077/2020\|EPI_ISL_749742\|202 | ···················· | ···················· | ···················· | ···················· | ···················· | ···················· |
| AA | 1096 hCoV-19/Denmark/DCGC-18079/2020\|EPI_ISL_749743\|202 | ···················· | ···················· | ···················· | ···················· | ···················· | ···················· |
| AA | 1097 hCoV-19/Denmark/DCGC-18080/2020\|EPI_ISL_749744\|202 | ···················· | ···················· | ···················· | ···················· | ···················· | ···················· |
| AA | 1098 hCoV-19/Denmark/DCGC-18082/2020\|EPI_ISL_749745\|202 | ···················· | ···················· | ···················· | ···················· | ···················· | ···················· |
| AA | 1099 hCoV-19/Denmark/DCGC-18083/2020\|EPI_ISL_749746\|202 | ···················· | ···················· | ···················· | ···················· | ···················· | ···················· |
| AA | 1100 hCoV-19/Denmark/DCGC-18084/2020\|EPI_ISL_749747\|202 | ···················· | ···················· | ···················· | ···················· | ···················· | ···················· |
| AA | 1101 hCoV-19/Denmark/DCGC-18088/2020\|EPI_ISL_749748\|202 | ···················· | ···················· | ···················· | ···················· | ···················· | ···················· |
| AA | 1102 hCoV-19/Denmark/DCGC-18089/2020\|EPI_ISL_749749\|202 | ···················· | ···················· | ···················· | ···················· | ······T············· | ···················· |
| AA | 1103 hCoV-19/Denmark/DCGC-18090/2020\|EPI_ISL_749750\|202 | ···················· | ···················· | ···················· | ···················· | ···················· | ···················· |
| AA | 1104 hCoV-19/Denmark/DCGC-18091/2020\|EPI_ISL_749751\|202 | ···················· | ···················· | ···················· | ···················· | ···················· | ···················· |
| AA | 1105 hCoV-19/Denmark/DCGC-18093/2020\|EPI_ISL_749752\|202 | ···················· | ···················· | ···················· | ···················· | ···················· | ···················· |
| AA | 1106 hCoV-19/Denmark/DCGC-18098/2020\|EPI_ISL_749753\|202 | ···················· | ···················· | ···················· | ···················· | ···················· | ···················· |
| AA | 1107 hCoV-19/Denmark/DCGC-18099/2020\|EPI_ISL_749754\|202 | ···················· | ···················· | ···················· | ···················· | ···················· | ···················· |
| AA | 1108 hCoV-19/Denmark/DCGC-18102/2020\|EPI_ISL_749755\|202 | ···················· | ···················· | ···················· | ···················· | ···················· | ···················· |
| AA | 1109 hCoV-19/Denmark/DCGC-18103/2020\|EPI_ISL_749756\|202 | ···················· | ···················· | ···················· | ···················· | ···················· | ···················· |
| AA | 1110 hCoV-19/Denmark/DCGC-18106/2020\|EPI_ISL_749757\|202 | ···················· | ···················· | ···················· | ···················· | ···················· | ···················· |
| AA | 1111 hCoV-19/Denmark/DCGC-18107/2020\|EPI_ISL_749758\|202 | ···················· | ···················· | ···················· | ···················· | ···················· | ···················· |
| AA | 1112 hCoV-19/Denmark/DCGC-18108/2020\|EPI_ISL_749759\|202 | ···················· | ···················· | ···················· | ···················· | ···················· | ···················· |
| AA | 1113 hCoV-19/Denmark/DCGC-16999/2020\|EPI_ISL_750700\|202 | ···················· | ·A·················· | ···················· | ···················· | ···················· | ···A················ |
| AA | 1114 hCoV-19/Denmark/DCGC-17000/2020\|EPI_ISL_750701\|202 | ···················· | ···················· | ···················· | ···················· | ···················· | ···················· |
| AA | 1115 hCoV-19/Denmark/DCGC-17004/2020\|EPI_ISL_750702\|202 | ···················· | ···················· | ···················· | ···················· | ···················· | ···················· |
| AA | 1116 hCoV-19/Denmark/DCGC-17005/2020\|EPI_ISL_750703\|202 | ···················· | ···················· | ···················· | ···················· | ···················· | ···················· |
| AA | 1117 hCoV-19/Denmark/DCGC-17007/2020\|EPI_ISL_750704\|202 | ···················· | ···················· | ···················· | ···················· | ···················· | ···················· |
| AA | 1118 hCoV-19/Denmark/DCGC-17008/2020\|EPI_ISL_750705\|202 | ···················· | ·A·················· | ···················· | ···················· | ···················· | ···················· |
| AA | 1119 hCoV-19/Denmark/DCGC-17009/2020\|EPI_ISL_750706\|202 | ···················· | ···················· | ···················· | ···················· | ···················· | ···················· |

AA 1120 hCoV-19/Denmark/DCGC-17011/2020|EPI_ISL_750707|202 ···················· ···················· ···················· ···················· ···················· ····················
AA 1121 hCoV-19/Denmark/DCGC-17012/2020|EPI_ISL_750708|202 ···················· ···················· ···················· ···················· ···················· ····················
AA 1122 hCoV-19/Denmark/DCGC-17013/2020|EPI_ISL_750709|202 ···················· ···················· ···················· ···················· ···················· ····················
AA 1123 hCoV-19/Denmark/DCGC-17014/2020|EPI_ISL_750710|202 ···················· ···················· ···················· ···················· ···················· ····················
AA 1124 hCoV-19/Denmark/DCGC-17015/2020|EPI_ISL_750711|202 ···················· ···················· ···················· ···················· ···················· ····················
AA 1125 hCoV-19/Denmark/DCGC-17016/2020|EPI_ISL_750712|202 ···················· ···················· ···················· ···················· ···················· ····················
AA 1126 hCoV-19/Denmark/DCGC-17018/2020|EPI_ISL_750713|202 ···················· ···················· ···················· ···················· ···················· ····················
AA 1127 hCoV-19/Denmark/DCGC-17019/2020|EPI_ISL_750714|202 ···················· ···················· ···················· ···················· ···················· ····················
AA 1128 hCoV-19/Denmark/DCGC-17020/2020|EPI_ISL_750715|202 ···················· ···················· ···················· ···················· ···················· ····················
AA 1129 hCoV-19/Denmark/DCGC-17021/2020|EPI_ISL_750716|202 ···················· ···················· ···················· ···················· ···················· ····················
AA 1130 hCoV-19/Denmark/DCGC-17023/2020|EPI_ISL_750717|202 ···················· ···················· ···················· ···················· ···················· ····················
AA 1131 hCoV-19/Denmark/DCGC-17026/2020|EPI_ISL_750718|202 ···················· ···················· ···················· ···················· ···················· ····················
AA 1132 hCoV-19/Denmark/DCGC-17027/2020|EPI_ISL_750719|202 ···················· ···················· ···················· ···················· ···················· ····················
AA 1133 hCoV-19/Denmark/DCGC-17028/2020|EPI_ISL_750720|202 ···················· ···················· ···················· ···················· ···················· ····················
AA 1134 hCoV-19/Denmark/DCGC-17033/2020|EPI_ISL_750721|202 ···················· ···················· ···················· ···················· ···················· ····················
AA 1135 hCoV-19/Denmark/DCGC-17036/2020|EPI_ISL_750722|202 ···················· ···················· ···················· ···················· ···················· ····················
AA 1136 hCoV-19/Denmark/DCGC-17037/2020|EPI_ISL_750723|202 ···················· ···················· ···················· ···················· ···················· ····················
AA 1137 hCoV-19/Denmark/DCGC-17041/2020|EPI_ISL_750724|202 ···················· ···················· ···················· ···················· ···················· ····················
AA 1138 hCoV-19/Denmark/DCGC-17042/2020|EPI_ISL_750725|202 ···················· ···················· ···················· ···················· ···················· ····················
AA 1139 hCoV-19/Denmark/DCGC-17043/2020|EPI_ISL_750726|202 ···················· ···················· ···················· ···················· ···················· ····················
AA 1140 hCoV-19/Denmark/DCGC-17045/2020|EPI_ISL_750727|202 ···················· ···················· ···················· ···················· ···················· ····················
AA 1141 hCoV-19/Denmark/DCGC-17046/2020|EPI_ISL_750728|202 ···················· ···················· ···················· ···················· ···················· ····················
AA 1142 hCoV-19/Denmark/DCGC-17047/2020|EPI_ISL_750729|202 ···················· ···················· ···················· ···················· ···················· ····················
AA 1143 hCoV-19/Denmark/DCGC-17048/2020|EPI_ISL_750730|202 ···················· ···················· ···················· ···················· ···················· ····················
AA 1144 hCoV-19/Denmark/DCGC-17050/2020|EPI_ISL_750731|202 ···················· ···················· ···················· ···················· ···················· ····················
AA 1145 hCoV-19/Denmark/DCGC-17052/2020|EPI_ISL_750732|202 ···················· ···················· ···················· ···················· ···················· ····················
AA 1146 hCoV-19/Denmark/DCGC-17054/2020|EPI_ISL_750733|202 ···················· ···················· ···················· ···················· ···················· ····················
AA 1147 hCoV-19/Denmark/DCGC-17056/2020|EPI_ISL_750734|202 ···················· ···················· ···················· ···················· ···················· ····················
AA 1148 hCoV-19/Denmark/DCGC-17057/2020|EPI_ISL_750735|202 ···················· ·A·················· ···················· ···················· ···················· ····················
AA 1149 hCoV-19/Denmark/DCGC-17059/2020|EPI_ISL_750736|202 ···················· ···················· ···················· ···················· ···················· ····················
AA 1150 hCoV-19/Denmark/DCGC-17060/2020|EPI_ISL_750737|202 ···················· ···················· ···················· ···················· ···················· ····················
AA 1151 hCoV-19/Denmark/DCGC-17061/2020|EPI_ISL_750738|202 ···················· ···················· ···················· ···················· ···················· ····················
AA 1152 hCoV-19/Denmark/DCGC-17063/2020|EPI_ISL_750739|202 ···················· ···················· ···················· ···················· ···················· ····················
AA 1153 hCoV-19/Denmark/DCGC-17064/2020|EPI_ISL_750740|202 ···················· ···················· ···················· ···················· ···················· ····················
AA 1154 hCoV-19/Russia/MOS-CRIE-13637504/2020|EPI_ISL_7542 ···················· ···················· ···················· ···················· ···················· ····················
AA 1155 hCoV-19/Russia/MOS-CRIE-13672647/2020|EPI_ISL_7542 ···················· ···················· ···················· ···················· ···················· ····················
AA 1156 hCoV-19/Russia/MOS-CRIE-13650805/2020|EPI_ISL_7542 ···················· ···················· ···················· ···················· ···················· ····················
AA 1157 hCoV-19/Russia/MOS-CRIE-13652490/2020|EPI_ISL_7542 ···················· ···················· ···················· ···················· ···················· ····················
AA 1158 hCoV-19/Russia/MOS-CRIE-13675724/2020|EPI_ISL_7542 ···················· ···················· ···················· ···················· ···················· ····················
AA 1159 hCoV-19/Russia/MOS-CRIE-13674129/2020|EPI_ISL_7542 ···················· ···················· ···················· ···················· ···················· ····················
AA 1160 hCoV-19/Russia/MOS-CRIE-13669866/2020|EPI_ISL_7542 ···················· ···················· ···················· ···················· ···················· ····················
AA 1161 hCoV-19/Russia/MOS-CRIE-13663504/2020|EPI_ISL_7542 ···················· ·A·················· ···················· ···················· ···················· ····················
AA 1162 hCoV-19/Russia/MOS-CRIE-13660226/2020|EPI_ISL_7542 ···················· ···················· ···················· ···················· ···················· ····················
AA 1163 hCoV-19/Russia/MOS-CRIE-13666589/2020|EPI_ISL_7542 ···················· ···················· ···················· ···················· ···················· ····················
AA 1164 hCoV-19/Russia/MOS-CRIE-13652759/2020|EPI_ISL_7542 ···················· ···················· ···················· ···················· ···················· ····················
AA 1165 hCoV-19/Russia/MOS-CRIE-13670319/2020|EPI_ISL_7542 ···················· ···················· ···················· ···················· ···················· ····················
AA 1166 hCoV-19/Russia/MOS-CRIE-13665359/2020|EPI_ISL_7542 ···················· ···················· ···················· ···················· ···················· ····················
AA 1167 hCoV-19/Russia/MOS-CRIE-13674490/2020|EPI_ISL_7542 ···················· ···················· ···················· ···················· ···················· ····················
AA 1168 hCoV-19/USA/WY-WPHL-20094903/2020|EPI_ISL_754215|2 ···················· ···················· ···················· ···················· ···················· ····················
AA 1169 hCoV-19/USA/WY-WPHL-20094982/2020|EPI_ISL_754216|2 ···················· ·A·················· ···················· ···················· ·NN················· ····················
AA 1170 hCoV-19/USA/WY-WPHL-20094985/2020|EPI_ISL_754217|2 ···················· ···················· ···················· ···················· ···················· ····················
AA 1171 hCoV-19/USA/WY-WPHL-20095054/2020|EPI_ISL_754218|2 ···················· ···················· ···················· ···················· ···················· ····················
AA 1172 hCoV-19/USA/WY-WPHL-20095066/2020|EPI_ISL_754219|2 ···················· ·A·················· ···················· ···················· ···················· ····················
AA 1173 hCoV-19/USA/WY-WPHL-20095104/2020|EPI_ISL_754220|2 ···················· ···················· ···················· ···················· ···················· ····················
AA 1174 hCoV-19/USA/WY-WPHL-20095131/2020|EPI_ISL_754221|2 ···················· ·A·················· ···················· ···················· ···················· ····················
AA 1175 hCoV-19/USA/WY-WPHL-20095188/2020|EPI_ISL_754223|2 ···················· ···················· ···················· ···················· ···················· ····················
AA 1176 hCoV-19/USA/WY-WPHL-20095212/2020|EPI_ISL_754224|2 ···················· ·A·················· ···················· ···················· ···················· ····················
AA 1177 hCoV-19/USA/WY-WPHL-20095283/2020|EPI_ISL_754225|2 ···················· ·A·················· ···················· ···················· ···················· ····················
AA 1178 hCoV-19/USA/WY-WPHL-20095317/2020|EPI_ISL_754226|2 ···················· ···················· ···················· ···················· ···················· ····················
AA 1179 hCoV-19/USA/WY-WPHL-20095417/2020|EPI_ISL_754227|2 ···················· ···················· ···················· ···················· ···················· ····················
AA 1180 hCoV-19/USA/WY-WPHL-20095509/2020|EPI_ISL_754228|2 ···················· ·A·················· ···················· ···················· ···················· ····················
AA 1181 hCoV-19/Belarus/HO-RII-MH11354S/2020|EPI_ISL_75423 ···················· ·A·················· ···················· ···················· ···················· ····················
AA 1182 hCoV-19/Belarus/MI-RII-MH11355S/2020|EPI_ISL_75423 ···················· ·A·················· ···················· ···················· ···················· ····················
AA 1184 hCoV-19/Romania/Bucuresti_407958/2020|EPI_ISL_7542 ···················· ·A·················· ···················· ···················· ···················· ····················
AA 1185 hCoV-19//Indonesia/JB-TFRIC19-R46646/2020|EPI_ISL_ ···················· ···················· ···················· ···················· ···················· ····················
AA 1186 hCoV-19/Indonesia/JB-TFRIC19-R50090/2020|EPI_ISL_7 ···················· ···A················ ···················· ···················· ···················· ·N··················
AA 1187 hCoV-19/England/204881101/2020|EPI_ISL_754287|2020 ···················· ···················· ···················· ···················· ···················· ····················
AA 1188 hCoV-19/England/204941555/2020|EPI_ISL_754290|2020 ···················· ···················· ···················· ···················· ···················· ····················
AA 1189 hCoV-19/England/204980818/2020|EPI_ISL_754291|2020 ···················· ···················· ···················· ···················· ···················· ····················
AA 1190 hCoV-19/England/204981914/2020|EPI_ISL_754292|2020 ···················· ···················· ···················· ···················· ···················· ····················
AA 1191 hCoV-19/England/204981916/2020|EPI_ISL_754293|2020 ···················· ···················· ···················· ···················· ···················· ····················
AA 1192 hCoV-19/England/204861491/2020|EPI_ISL_754294|2020 ···················· ···················· ···················· ···················· ···················· ····················
AA 1193 hCoV-19/England/204881368/2020|EPI_ISL_754295|2020 ···················· ···················· ···················· ···················· ···················· ····················
AA 1194 hCoV-19/England/204881370/2020|EPI_ISL_754296|2020 ···················· ···················· ···················· ···················· ···················· ····················
AA 1195 hCoV-19/England/204881371/2020|EPI_ISL_754297|2020 ···················· ···················· ···················· ···················· ······T············· ····················
AA 1196 hCoV-19/England/204941330/2020|EPI_ISL_754298|2020 ···················· ···················· ···················· ···················· ···················· ····················
AA 1197 hCoV-19/England/204941388/2020|EPI_ISL_754299|2020 ···················· ···················· ···················· ···················· ···················· ····················
AA 1198 hCoV-19/USA/ME-HETL-J0409/2020|EPI_ISL_755517|2020 ···················· ···················· ···················· ···················· ···················· ····················
AA 1199 hCoV-19/USA/ME-HETL-J0410/2020|EPI_ISL_755518|2020 ···················· ···················· ···················· ···················· ···················· ····················
AA 1200 hCoV-19/USA/ME-HETL-J0411/2020|EPI_ISL_755519|2020 ···················· ···················· ···················· ···················· ···················· ····················

| | | | | | | | | | |
|---|---|---|---|---|---|---|---|---|---|
| AA | 1201 | hCoV-19/USA/ME-HETL-J0414/2020\|EPI_ISL_755520\|2020 | ···················· | ···················· | ···················· | ···················· | ···················· | ···················· | |
| AA | 1202 | hCoV-19/USA/ME-HETL-J0418/2020\|EPI_ISL_755522\|2020 | ··················T· | ·A·················· | ···················· | ···················· | ···················· | ···················· | 1 |
| AA | 1203 | hCoV-19/USA/ME-HETL-J0419/2020\|EPI_ISL_755523\|2020 | ···················· | ···················· | ···················· | ···················· | ···················· | ···················· | |
| AA | 1204 | hCoV-19/USA/ME-HETL-J0421/2020\|EPI_ISL_755524\|2020 | ··················T· | ·A·················· | ···················· | ···················· | ···················· | ···················· | 1 |
| AA | 1205 | hCoV-19/USA/ME-HETL-J0422/2020\|EPI_ISL_755525\|2020 | ··················T· | ·A·················· | ···················· | ···················· | ···················· | ···················· | 1 |
| AA | 1206 | hCoV-19/USA/ME-HETL-J0423/2020\|EPI_ISL_755526\|2020 | ···················· | ···················· | ···················· | ···················· | ···················· | ···················· | |
| AA | 1207 | hCoV-19/USA/ME-HETL-J0425/2020\|EPI_ISL_755527\|2020 | ···················· | ···················· | ···················· | ···················· | ···················· | ···················· | |
| AA | 1209 | hCoV-19/USA/ME-HETL-J0427/2020\|EPI_ISL_755529\|2020 | ···················· | ···················· | ···················· | ···················· | ···················· | ···················· | |
| AA | 1210 | hCoV-19/USA/ME-HETL-J0428/2020\|EPI_ISL_755530\|2020 | ···················· | ···················· | ···················· | ···················· | ···················· | ···················· | |
| AA | 1211 | hCoV-19/USA/ME-HETL-J0430/2020\|EPI_ISL_755531\|2020 | ···················· | ···················· | ···················· | ···················· | ···················· | ···················· | |
| AA | 1213 | hCoV-19/USA/ME-HETL-J0433/2020\|EPI_ISL_755533\|2020 | ···················· | ···················· | ···················· | ···················· | ···················· | ···················· | |
| AA | 1214 | hCoV-19/USA/ME-HETL-J0434/2020\|EPI_ISL_755534\|2020 | ···················· | ···················· | ···················· | ···················· | ···················· | ···················· | |
| AA | 1215 | hCoV-19/USA/ME-HETL-J0435/2020\|EPI_ISL_755535\|2020 | ···················· | ···················· | ···················· | ···················· | ···················· | ···················· | |
| AA | 1216 | hCoV-19/USA/ME-HETL-J0436/2020\|EPI_ISL_755536\|2020 | ···················· | ···················· | ···················· | ···················· | ···················· | ···················· | |
| AA | 1217 | hCoV-19/USA/ME-HETL-J0437/2020\|EPI_ISL_755537\|2020 | ···················· | ···················· | ···················· | ···················· | ···················· | ···················· | |
| AA | 1218 | hCoV-19/USA/ME-HETL-J0438/2020\|EPI_ISL_755538\|2020 | ···················· | ···················· | ···················· | ···················· | ···················· | ···················· | |
| AA | 1219 | hCoV-19/USA/ME-HETL-J0440/2020\|EPI_ISL_755539\|2020 | ···················· | ···················· | ···················· | ···················· | ···················· | ···················· | |
| AA | 1220 | hCoV-19/USA/ME-HETL-J0441/2020\|EPI_ISL_755540\|2020 | ···················· | ···················· | ···················· | ···················· | ···················· | ···················· | |
| AA | 1221 | hCoV-19/USA/ME-HETL-J0442/2020\|EPI_ISL_755541\|2020 | ···················· | ···················· | ···················· | ···················· | ···················· | ···················· | |
| AA | 1222 | hCoV-19/USA/ME-HETL-J0445/2020\|EPI_ISL_755543\|2020 | ···················· | ···················· | ···················· | ···················· | ···················· | ···················· | |
| AA | 1223 | hCoV-19/USA/ME-HETL-J0446/2020\|EPI_ISL_755544\|2020 | ···················· | ···················· | ···················· | ···················· | ···················· | ···················· | |
| AA | 1224 | hCoV-19/USA/ME-HETL-J0447/2020\|EPI_ISL_755545\|2020 | ···················· | ···················· | ···················· | ···················· | ···················· | ···················· | |
| AA | 1225 | hCoV-19/USA/ME-HETL-J0448/2020\|EPI_ISL_755546\|2020 | ···················· | ···················· | ···················· | ···················· | ···················· | ···················· | |
| AA | 1226 | hCoV-19/USA/ME-HETL-J0449/2020\|EPI_ISL_755547\|2020 | ···················· | ···················· | ···················· | ···················· | ···················· | ···················· | |
| AA | 1227 | hCoV-19/USA/ME-HETL-J0451/2020\|EPI_ISL_755548\|2020 | ···················· | ·A·················· | ···················· | ···················· | ···················· | ···················· | |
| AA | 1228 | hCoV-19/USA/ME-HETL-J0453/2020\|EPI_ISL_755549\|2020 | ···················· | ···················· | ···················· | ···················· | ···················· | ···················· | |
| AA | 1229 | hCoV-19/USA/ME-HETL-J0458/2020\|EPI_ISL_755551\|2020 | ···················· | ···················· | ···················· | ···················· | ···················· | ···················· | |
| AA | 1230 | hCoV-19/USA/ME-HETL-J0461/2020\|EPI_ISL_755553\|2020 | ···················· | ···················· | ···················· | ···················· | ···················· | ···················· | |
| AA | 1231 | hCoV-19/USA/ME-HETL-J0464/2020\|EPI_ISL_755556\|2020 | ···················· | ···················· | ···················· | ···················· | ···················· | ···················· | |
| AA | 1232 | hCoV-19/USA/ME-HETL-J0466/2020\|EPI_ISL_755557\|2020 | ···················· | ·A·················· | ···················· | ···················· | ···················· | ···················· | |
| AA | 1233 | hCoV-19/USA/ME-HETL-J0467/2020\|EPI_ISL_755558\|2020 | ···················· | ···················· | ···················· | ···················· | ···················· | ···················· | |
| AA | 1234 | hCoV-19/USA/ME-HETL-J0468/2020\|EPI_ISL_755559\|2020 | ···················· | ···················· | ···················· | ···················· | ···················· | ···················· | |
| AA | 1235 | hCoV-19/USA/ME-HETL-J0469/2020\|EPI_ISL_755560\|2020 | ···················· | ···················· | ···················· | ···················· | ···················· | ···················· | |
| AA | 1236 | hCoV-19/USA/ME-HETL-J0470/2020\|EPI_ISL_755561\|2020 | ···················· | ···················· | ···················· | ···················· | ···················· | ···················· | |
| AA | 1237 | hCoV-19/USA/ME-HETL-J0471/2020\|EPI_ISL_755562\|2020 | ···················· | ···················· | ···················· | ···················· | ···················· | ···················· | |
| AA | 1238 | hCoV-19/USA/ME-HETL-J0472/2020\|EPI_ISL_755563\|2020 | ···················· | ·A·················· | ···················· | ···················· | ···················· | ···················· | |
| AA | 1239 | hCoV-19/USA/ME-HETL-J0473/2020\|EPI_ISL_755564\|2020 | ···················· | ···················· | ···················· | ···················· | ···················· | ···················· | |
| AA | 1240 | hCoV-19/USA/ME-HETL-J0474/2020\|EPI_ISL_755565\|2020 | ···················· | ···················· | ···················· | ···················· | ···················· | ···················· | |
| AA | 1241 | hCoV-19/USA/ME-HETL-J0477/2020\|EPI_ISL_755566\|2020 | ···················· | ···················· | ···················· | ···················· | ···················· | ···················· | |
| AA | 1242 | hCoV-19/USA/ME-HETL-J0480/2020\|EPI_ISL_755567\|2020 | ···················· | ·A·················· | ···················· | ···················· | ···················· | ···················· | |
| AA | 1243 | hCoV-19/Australia/SAP598/2020\|EPI_ISL_755568\|2020- | ···················· | ·A·················· | ···················· | ···················· | ···················· | ···················· | |
| AA | 1244 | hCoV-19/Australia/SAP600/2020\|EPI_ISL_755570\|2020- | ···················· | ·A·················· | ···················· | ···················· | ···················· | ···················· | |
| AA | 1245 | hCoV-19/Australia/SAP601/2020\|EPI_ISL_755571\|2020- | ···················· | ···················· | ···················· | ···················· | ···················· | ···················· | |
| AA | 1246 | hCoV-19/Italy/ABR-CAST-CH360/2020\|EPI_ISL_755572\|2 | ···················· | ···················· | ···················· | ···················· | ···················· | ···················· | |
| AA | 1247 | hCoV-19/Italy/ABR-CAST-CB543/2020\|EPI_ISL_755573\|2 | ···················· | ···················· | ···················· | ···················· | ···················· | ···················· | |
| AA | 1248 | hCoV-19/Italy/ABR-CAST-CL106/2020\|EPI_ISL_755574\|2 | ···················· | ···················· | ···················· | ···················· | ···················· | ···················· | |
| AA | 1249 | hCoV-19/India/UN-THSTI-BL2010D/2020\|EPI_ISL_755587 | ···················· | ···················· | ···················· | ···················· | ···················· | ···················· | |
| AA | 1250 | hCoV-19/USA/MA-CDC-STM-P003/2020\|EPI_ISL_755590\|20 | ···················· | ···················· | ···················· | ···················· | ···················· | ···················· | |
| AA | 1251 | hCoV-19/USA/PA-CDC-STM-P010/2020\|EPI_ISL_755591\|20 | ···················· | ···················· | ···················· | ···················· | ···················· | ···················· | |
| AA | 1252 | hCoV-19/USA/FL-CDC-STM-P058/2020\|EPI_ISL_755592\|20 | ···················· | ···················· | ···················· | ···················· | ···················· | ···················· | |
| AA | 1253 | hCoV-19/USA/CA-CDC-STM-P002/2020\|EPI_ISL_755597\|20 | ···················· | ···················· | ···················· | ···················· | ···················· | ···················· | |
| AA | 1254 | hCoV-19/USA/FL-CDC-STM-P004/2020\|EPI_ISL_755598\|20 | ···················· | ···················· | ···················· | ···················· | ···················· | ···················· | |
| AA | 1255 | hCoV-19/USA/FL-CDC-STM-P005/2020\|EPI_ISL_755599\|20 | ···················· | ···················· | ···················· | ···················· | ···················· | ···················· | |
| AA | 1256 | hCoV-19/England/QEUH-C7C7B3/2020\|EPI_ISL_756850\|20 | ···················· | ···················· | ···················· | ···················· | ···················· | ···················· | |
| AA | 1257 | hCoV-19/England/MILK-C78456/2020\|EPI_ISL_756857\|20 | ···················· | ···················· | ···················· | ···················· | ···················· | ···················· | |
| AA | 1258 | hCoV-19/England/MILK-C7B778/2020\|EPI_ISL_756860\|20 | ···················· | ···················· | ···················· | ···················· | ···················· | ···················· | |
| AA | 1259 | hCoV-19/England/MILK-C7B3CF/2020\|EPI_ISL_756863\|20 | ···················· | ···················· | ···················· | ···················· | ···················· | ···················· | |
| AA | 1260 | hCoV-19/England/MILK-C7B75A/2020\|EPI_ISL_756865\|20 | ···················· | ···················· | ···················· | ···················· | ···················· | ···················· | |
| AA | 1261 | hCoV-19/England/MILK-C7B918/2020\|EPI_ISL_756868\|20 | ···················· | ···················· | ···················· | ···················· | ···················· | ···················· | |
| AA | 1262 | hCoV-19/England/MILK-C78508/2020\|EPI_ISL_756871\|20 | ··················T· | ···················· | ···················· | ···················· | ···················· | ···················· | 1 |
| AA | 1263 | hCoV-19/England/MILK-C796C8/2020\|EPI_ISL_756872\|20 | ···················· | ···················· | ···················· | ···················· | ···················· | ···················· | |
| AA | 1264 | hCoV-19/England/MILK-C7B365/2020\|EPI_ISL_756873\|20 | ···················· | ···················· | ···················· | ···················· | ···················· | ···················· | |
| AA | 1265 | hCoV-19/England/MILK-C7B550/2020\|EPI_ISL_756875\|20 | ···················· | ···················· | ···················· | ···················· | ···················· | ···················· | |
| AA | 1266 | hCoV-19/England/QEUH-C801BE/2020\|EPI_ISL_756879\|20 | ···················· | ···················· | ···················· | ···················· | ···················· | ···················· | |
| AA | 1267 | hCoV-19/England/MILK-C7B435/2020\|EPI_ISL_756880\|20 | ···················· | ···················· | ···················· | ···················· | ······T············· | ···················· | |
| AA | 1268 | hCoV-19/England/QEUH-C7D7B2/2020\|EPI_ISL_756889\|20 | ···················· | ···················· | ···················· | ···················· | ···················· | ···················· | |
| AA | 1269 | hCoV-19/England/QEUH-C7C294/2020\|EPI_ISL_756893\|20 | ···················· | ···················· | ···················· | ···················· | ···················· | ···················· | |
| AA | 1270 | hCoV-19/England/QEUH-C7D8CE/2020\|EPI_ISL_756895\|20 | ···················· | ···················· | ···················· | ···················· | ···················· | ···················· | |
| AA | 1271 | hCoV-19/England/QEUH-C7FEF7/2020\|EPI_ISL_756897\|20 | ···················· | ···················· | ···················· | ···················· | ···················· | ···················· | |
| AA | 1272 | hCoV-19/England/MILK-C7822F/2020\|EPI_ISL_756899\|20 | ···················· | ···················· | ···················· | ···················· | ···················· | ···················· | |
| AA | 1273 | hCoV-19/Denmark/DCGC-21048/2020\|EPI_ISL_759053\|202 | ···················· | ·A·················· | ···················· | ···················· | ···················· | ···A················ | |
| AA | 1274 | hCoV-19/Denmark/DCGC-21205/2020\|EPI_ISL_759056\|202 | ···················· | ···················· | ···················· | ···················· | ···················· | ···················· | |
| AA | 1275 | hCoV-19/Denmark/DCGC-21248/2020\|EPI_ISL_759057\|202 | ···················· | ·A·················· | ···················· | ···················· | ···················· | ···A················ | |
| AA | 1276 | hCoV-19/Denmark/DCGC-21260/2020\|EPI_ISL_759058\|202 | ···················· | ···················· | ···················· | ···················· | ···················· | ···················· | |
| AA | 1277 | hCoV-19/Denmark/DCGC-21288/2020\|EPI_ISL_759059\|202 | ···················· | ·A·················· | ···················· | ···················· | ···················· | ···A················ | |
| AA | 1278 | hCoV-19/Denmark/DCGC-20988/2020\|EPI_ISL_759065\|202 | ···················· | ···················· | ···················· | ···················· | ···················· | ···················· | |
| AA | 1279 | hCoV-19/Denmark/DCGC-20954/2020\|EPI_ISL_759075\|202 | ···················· | ···················· | ···················· | ···················· | ···················· | ···················· | |
| AA | 1280 | hCoV-19/Denmark/DCGC-21313/2020\|EPI_ISL_759076\|202 | ···················· | ···················· | ···················· | ···················· | ···················· | ···················· | |
| AA | 1281 | hCoV-19/USA/WA-UW-45052/2020\|EPI_ISL_747195\|2020-1 | ···················· | ···················· | ···················· | ···················· | ···················· | ···················· | |
| AA | 1282 | hCoV-19/USA/WA-UW-45058/2020\|EPI_ISL_747196\|2020-1 | ···················· | ···················· | ···················· | ···················· | ···················· | ···················· | |

| | | | | | | | | |
|---|---|---|---|---|---|---|---|---|
| AA | 1283 | hCoV-19/USA/WA-UW-45059/2020\|EPI_ISL_747197\|2020-1 | ···················· | ···················· | ····················· | ···················· | ···················· | ···················· |
| AA | 1284 | hCoV-19/USA/WA-UW-45060/2020\|EPI_ISL_747198\|2020-1 | ···················· | ···················· | ····················· | ···················· | ···················· | ···················· |
| AA | 1285 | hCoV-19/USA/WA-UW-45061/2020\|EPI_ISL_747199\|2020-1 | ···················· | ···················· | ····················· | ···················· | ···················· | ···················· |
| AA | 1286 | hCoV-19/Denmark/DCGC-19272/2020\|EPI_ISL_748430\|202 | ···················· | ···················· | ····················· | ···················· | ···················· | ···················· |
| AA | 1287 | hCoV-19/Denmark/DCGC-19282/2020\|EPI_ISL_748431\|202 | ···················· | ···················· | ····················· | ···················· | ···················· | ···················· |
| AA | 1288 | hCoV-19/Denmark/DCGC-19291/2020\|EPI_ISL_748432\|202 | ···················· | ···················· | ····················· | ···················· | ···················· | ···················· |
| AA | 1289 | hCoV-19/Denmark/DCGC-19294/2020\|EPI_ISL_748433\|202 | ···················· | ···················· | ····················· | ···················· | ···················· | ···················· |
| AA | 1290 | hCoV-19/Denmark/DCGC-19305/2020\|EPI_ISL_748434\|202 | ···················· | ···················· | ····················· | ···················· | ···················· | ···················· |
| AA | 1291 | hCoV-19/Denmark/DCGC-19308/2020\|EPI_ISL_748435\|202 | ···················· | ···················· | ····················· | ···················· | ···················· | ···················· |
| AA | 1292 | hCoV-19/Denmark/DCGC-19309/2020\|EPI_ISL_748436\|202 | ···················· | ···················· | ····················· | ···················· | ···················· | ···················· |
| AA | 1293 | hCoV-19/Denmark/DCGC-19311/2020\|EPI_ISL_748437\|202 | ···················· | ···················· | ····················· | ···················· | ···················· | ···················· |
| AA | 1294 | hCoV-19/Denmark/DCGC-19314/2020\|EPI_ISL_748438\|202 | ···················· | ···················· | ····················· | ···················· | ···················· | ···················· |
| AA | 1295 | hCoV-19/Denmark/DCGC-19321/2020\|EPI_ISL_748439\|202 | ···················· | ···················· | ····················· | ···················· | ···················· | ···················· |
| AA | 1296 | hCoV-19/Denmark/DCGC-19353/2020\|EPI_ISL_748440\|202 | ···················· | ···················· | ····················· | ···················· | ···················· | ···················· |
| AA | 1297 | hCoV-19/Denmark/DCGC-19354/2020\|EPI_ISL_748441\|202 | ···················· | ···················· | ····················· | ···················· | ···················· | ···················· |
| AA | 1298 | hCoV-19/Denmark/DCGC-19363/2020\|EPI_ISL_748442\|202 | ···················· | ···················· | ····················· | ···················· | ···················· | ···················· |
| AA | 1299 | hCoV-19/Denmark/DCGC-19388/2020\|EPI_ISL_748443\|202 | ···················· | ···················· | ····················· | ···················· | ···················· | ···················· |
| AA | 1300 | hCoV-19/Denmark/DCGC-19414/2020\|EPI_ISL_748444\|202 | ···················· | ···················· | ····················· | ···················· | ···················· | ···················· |
| AA | 1301 | hCoV-19/Denmark/DCGC-19417/2020\|EPI_ISL_748445\|202 | ···················· | ···················· | ····················· | ···················· | ···················· | ···················· |
| AA | 1302 | hCoV-19/Denmark/DCGC-19418/2020\|EPI_ISL_748446\|202 | ···················· | ···················· | ····················· | ···················· | ···················· | ···················· |
| AA | 1303 | hCoV-19/Denmark/DCGC-19422/2020\|EPI_ISL_748447\|202 | ···················· | ···················· | ····················· | ···················· | ···················· | ···················· |
| AA | 1304 | hCoV-19/Denmark/DCGC-19427/2020\|EPI_ISL_748448\|202 | ···················· | ···················· | ····················· | ···················· | ···················· | ···················· |
| AA | 1305 | hCoV-19/Denmark/DCGC-19461/2020\|EPI_ISL_748449\|202 | ···················· | ···················· | ····················· | ···················· | ···················· | ···················· |
| AA | 1306 | hCoV-19/Denmark/DCGC-19463/2020\|EPI_ISL_748450\|202 | ···················· | ···················· | ····················· | ···················· | ···················· | ···················· |
| AA | 1307 | hCoV-19/Denmark/DCGC-19483/2020\|EPI_ISL_748451\|202 | ···················· | ···················· | ····················· | ···················· | ···················· | ···················· |
| AA | 1308 | hCoV-19/Denmark/DCGC-19486/2020\|EPI_ISL_748452\|202 | ···················· | ···················· | ····················· | ···················· | ···················· | ···················· |
| AA | 1309 | hCoV-19/Denmark/DCGC-19500/2020\|EPI_ISL_748453\|202 | ···················· | ···················· | ····················· | ···················· | ···················· | ···················· |
| AA | 1310 | hCoV-19/Denmark/DCGC-19539/2020\|EPI_ISL_748454\|202 | ···················· | ···················· | ····················· | ···················· | ···················· | ···················· |
| AA | 1311 | hCoV-19/Denmark/DCGC-19546/2020\|EPI_ISL_748455\|202 | ···················· | ···················· | ····················· | ···················· | ···················· | ···················· |
| AA | 1312 | hCoV-19/Denmark/DCGC-19573/2020\|EPI_ISL_748456\|202 | ···················· | ···················· | ····················· | ···················· | ···················· | ···················· |
| AA | 1313 | hCoV-19/Denmark/DCGC-19575/2020\|EPI_ISL_748457\|202 | ···················· | ···················· | ····················· | ···················· | ···················· | ···················· |
| AA | 1314 | hCoV-19/Denmark/DCGC-19589/2020\|EPI_ISL_748458\|202 | ···················· | ···················· | ····················· | ···················· | ···················· | ···················· |
| AA | 1315 | hCoV-19/Denmark/DCGC-19621/2020\|EPI_ISL_748459\|202 | ···················· | ···················· | ····················· | ···················· | ···················· | ···················· |
| AA | 1316 | hCoV-19/Denmark/DCGC-19639/2020\|EPI_ISL_748460\|202 | ···················· | ···················· | ····················· | ···················· | ···················· | ···················· |
| AA | 1317 | hCoV-19/Denmark/DCGC-19642/2020\|EPI_ISL_748461\|202 | ···················· | ···················· | ····················· | ···················· | ···················· | ···················· |
| AA | 1318 | hCoV-19/Denmark/DCGC-19650/2020\|EPI_ISL_748462\|202 | ···················· | ···················· | ····················· | ···················· | ···················· | ···················· |
| AA | 1319 | hCoV-19/Denmark/DCGC-19663/2020\|EPI_ISL_748463\|202 | ···················· | ···················· | ····················· | ···················· | ···················· | ···················· |
| AA | 1320 | hCoV-19/Denmark/DCGC-19688/2020\|EPI_ISL_748464\|202 | ···················· | ···················· | ····················· | ···················· | ···················· | ···················· |
| AA | 1321 | hCoV-19/Denmark/DCGC-19696/2020\|EPI_ISL_748465\|202 | ···················· | ···················· | ····················· | ···················· | ···················· | ···················· |
| AA | 1322 | hCoV-19/Denmark/DCGC-19704/2020\|EPI_ISL_748466\|202 | ···················· | ···················· | ····················· | ···················· | ···················· | ···················· |
| AA | 1323 | hCoV-19/Denmark/DCGC-19721/2020\|EPI_ISL_748467\|202 | ···················· | ···················· | ····················· | ···················· | ···················· | ···················· |
| AA | 1324 | hCoV-19/Denmark/DCGC-19724/2020\|EPI_ISL_748468\|202 | ···················· | ···················· | ····················· | ···················· | ···················· | ···················· |
| AA | 1325 | hCoV-19/Denmark/DCGC-19741/2020\|EPI_ISL_748469\|202 | ···················· | ···················· | ····················· | ···················· | ···················· | ···················· |
| AA | 1326 | hCoV-19/Denmark/DCGC-19747/2020\|EPI_ISL_748470\|202 | ···················· | ···················· | ····················· | ···················· | ···················· | ···················· |
| AA | 1327 | hCoV-19/Denmark/DCGC-19748/2020\|EPI_ISL_748471\|202 | ···················· | ···················· | ····················· | ···················· | ···················· | ···················· |
| AA | 1328 | hCoV-19/Denmark/DCGC-18797/2020\|EPI_ISL_748484\|202 | ···················· | ·A·················· | ····················· | ···················· | ···················· | ··A················· |
| AA | 1329 | hCoV-19/Denmark/DCGC-19009/2020\|EPI_ISL_748485\|202 | ···················· | ···················· | ····················· | ···················· | ···················· | ···················· |
| AA | 1330 | hCoV-19/Denmark/DCGC-19334/2020\|EPI_ISL_748486\|202 | ···················· | ···················· | ····················· | ···················· | ···················· | ···················· |
| AA | 1331 | hCoV-19/Denmark/DCGC-19576/2020\|EPI_ISL_748487\|202 | ···················· | ···················· | ····················· | ···················· | ···················· | ···················· |
| AA | 1332 | hCoV-19/Denmark/DCGC-19598/2020\|EPI_ISL_748488\|202 | ···················· | ·A·················· | ····················· | ···················· | ···················· | ··A················· |
| AA | 1333 | hCoV-19/Denmark/DCGC-19605/2020\|EPI_ISL_748489\|202 | ···················· | ···················· | ····················· | ···················· | ···················· | ···················· |
| AA | 1334 | hCoV-19/Denmark/DCGC-19635/2020\|EPI_ISL_748490\|202 | ···················· | ···················· | ····················· | ···················· | ···················· | ···················· |
| AA | 1335 | hCoV-19/Denmark/DCGC-19654/2020\|EPI_ISL_748491\|202 | ···················· | ···················· | ····················· | ···················· | ···················· | ···················· |
| AA | 1336 | hCoV-19/Denmark/DCGC-19683/2020\|EPI_ISL_748492\|202 | ···················· | ···················· | ····················· | ···················· | ···················· | ···················· |
| AA | 1337 | hCoV-19/Denmark/DCGC-18041/2020\|EPI_ISL_749719\|202 | ···················· | ···················· | ····················· | ···················· | ···················· | ···················· |
| AA | 1338 | hCoV-19/Denmark/DCGC-18109/2020\|EPI_ISL_749760\|202 | ···················· | ···················· | ····················· | ···················· | ···················· | ···················· |
| AA | 1339 | hCoV-19/Denmark/DCGC-18110/2020\|EPI_ISL_749761\|202 | ···················· | ·············A······ | ····················· | ···················· | ···················· | ···················· |
| AA | 1340 | hCoV-19/Denmark/DCGC-18112/2020\|EPI_ISL_749762\|202 | ···················· | ···················· | ····················· | ···················· | ···················· | ···················· |
| AA | 1341 | hCoV-19/Denmark/DCGC-18114/2020\|EPI_ISL_749763\|202 | ···················· | ···················· | ····················· | ···················· | ···················· | ···················· |
| AA | 1342 | hCoV-19/Denmark/DCGC-18115/2020\|EPI_ISL_749764\|202 | ···················· | ···················· | ····················· | ···················· | ···················· | ···················· |
| AA | 1343 | hCoV-19/Denmark/DCGC-18117/2020\|EPI_ISL_749765\|202 | ················T··· | ···················· | ····················· | ···················· | ···················· | ···················· |
| AA | 1344 | hCoV-19/Denmark/DCGC-18118/2020\|EPI_ISL_749766\|202 | ···················· | ···················· | ····················· | ···················· | ···················· | ···················· |
| AA | 1345 | hCoV-19/Denmark/DCGC-18120/2020\|EPI_ISL_749767\|202 | ···················· | ···················· | ····················· | ···················· | ···················· | ···················· |
| AA | 1346 | hCoV-19/Denmark/DCGC-18122/2020\|EPI_ISL_749768\|202 | ···················· | ···················· | ····················· | ···················· | ···················· | ···················· |
| AA | 1347 | hCoV-19/Denmark/DCGC-18124/2020\|EPI_ISL_749769\|202 | ···················· | ···················· | ····················· | ···················· | ···················· | ···················· |
| AA | 1348 | hCoV-19/Denmark/DCGC-18125/2020\|EPI_ISL_749770\|202 | ···················· | ········C··········· | ····················· | ···················· | ···················· | ···················· |
| AA | 1349 | hCoV-19/Denmark/DCGC-18129/2020\|EPI_ISL_749771\|202 | ···················· | ···················· | ····················· | ···················· | ···················· | ···················· |
| AA | 1350 | hCoV-19/Denmark/DCGC-18133/2020\|EPI_ISL_749772\|202 | ···················· | ···················· | ····················· | ···················· | ···················· | ···················· |
| AA | 1351 | hCoV-19/Denmark/DCGC-18136/2020\|EPI_ISL_749773\|202 | ···················· | ···················· | ····················· | ···················· | ···················· | ···················· |
| AA | 1352 | hCoV-19/Denmark/DCGC-18137/2020\|EPI_ISL_749774\|202 | ···················· | ···················· | ····················· | ···················· | ···················· | ···················· |
| AA | 1353 | hCoV-19/Denmark/DCGC-18139/2020\|EPI_ISL_749775\|202 | ···················· | ···················· | ····················· | ···················· | ···················· | ···················· |
| AA | 1354 | hCoV-19/Denmark/DCGC-18140/2020\|EPI_ISL_749776\|202 | ···················· | ···················· | ····················· | ···················· | ···················· | ···················· |
| AA | 1355 | hCoV-19/Denmark/DCGC-18142/2020\|EPI_ISL_749777\|202 | ···················· | ···················· | ····················· | ···················· | ···················· | ···················· |
| AA | 1356 | hCoV-19/Denmark/DCGC-18144/2020\|EPI_ISL_749778\|202 | ···················· | ···················· | ····················· | ···················· | ···················· | ···················· |
| AA | 1357 | hCoV-19/Denmark/DCGC-18145/2020\|EPI_ISL_749779\|202 | ···················· | ···················· | ····················· | ···················· | ···················· | ···················· |
| AA | 1358 | hCoV-19/Denmark/DCGC-18147/2020\|EPI_ISL_749780\|202 | ···················· | ···················· | ····················· | ···················· | ···················· | ···················· |
| AA | 1359 | hCoV-19/Denmark/DCGC-18148/2020\|EPI_ISL_749781\|202 | ···················· | ···················· | ····················· | ···················· | ···················· | ···················· |
| AA | 1360 | hCoV-19/Denmark/DCGC-18150/2020\|EPI_ISL_749782\|202 | ···················· | ···················· | ····················· | ···················· | ···················· | ···················· |
| AA | 1361 | hCoV-19/Denmark/DCGC-18152/2020\|EPI_ISL_749783\|202 | ···················· | ···················· | ····················· | ···················· | ···················· | ···················· |
| AA | 1362 | hCoV-19/Denmark/DCGC-18153/2020\|EPI_ISL_749784\|202 | ···················· | ···················· | ····················· | ···················· | ···················· | ···················· |

| | | | | | | | | |
|---|---|---|---|---|---|---|---|---|
| AA | 1363 | hCoV-19/Denmark/DCGC-18155/2020\|EPI_ISL_749785\|202 | ···················· | ···················· | ····················· | ···················· | ···················· | ···················· |
| AA | 1364 | hCoV-19/Denmark/DCGC-18157/2020\|EPI_ISL_749786\|202 | ···················· | ···················· | ····················· | ···················· | ···················· | ···················· |
| AA | 1365 | hCoV-19/Denmark/DCGC-18162/2020\|EPI_ISL_749787\|202 | ···················· | ···················· | ····················· | ···················· | ···················· | ···················· |
| AA | 1366 | hCoV-19/Denmark/DCGC-18163/2020\|EPI_ISL_749788\|202 | ···················· | ···················· | ····················· | ···················· | ···················· | ···················· |
| AA | 1367 | hCoV-19/Denmark/DCGC-18164/2020\|EPI_ISL_749789\|202 | ···················· | ···················· | ····················· | ···················· | ···················· | ···················· |
| AA | 1368 | hCoV-19/Denmark/DCGC-18165/2020\|EPI_ISL_749790\|202 | ···················· | ···················· | ····················· | ···················· | ···················· | ···················· |
| AA | 1369 | hCoV-19/Denmark/DCGC-18167/2020\|EPI_ISL_749791\|202 | ···················· | ···················· | ····················· | ···················· | ···················· | ···················· |
| AA | 1370 | hCoV-19/Denmark/DCGC-18169/2020\|EPI_ISL_749792\|202 | ···················· | ···················· | ····················· | ···················· | ···················· | ···················· |
| AA | 1371 | hCoV-19/Denmark/DCGC-18170/2020\|EPI_ISL_749793\|202 | ···················· | ···················· | ····················· | ···················· | ···················· | ···················· |
| AA | 1372 | hCoV-19/Denmark/DCGC-18171/2020\|EPI_ISL_749794\|202 | ···················· | ···················· | ····················· | ···················· | ···················· | ···················· |
| AA | 1373 | hCoV-19/Denmark/DCGC-18173/2020\|EPI_ISL_749795\|202 | ···················· | ···················· | ····················· | ···················· | ···················· | ···················· |
| AA | 1374 | hCoV-19/Denmark/DCGC-18177/2020\|EPI_ISL_749796\|202 | ···················· | ···················· | ····················· | ···················· | ···················· | ···················· |
| AA | 1375 | hCoV-19/Denmark/DCGC-18179/2020\|EPI_ISL_749797\|202 | ···················· | ···················· | ····················· | ···················· | ···················· | ···················· |
| AA | 1376 | hCoV-19/Denmark/DCGC-18180/2020\|EPI_ISL_749798\|202 | ···················· | ···················· | ····················· | ···················· | ···················· | ···················· |
| AA | 1377 | hCoV-19/Denmark/DCGC-18182/2020\|EPI_ISL_749799\|202 | ···················· | ···················· | ····················· | ···················· | ···················· | ···················· |
| AA | 1378 | hCoV-19/Denmark/DCGC-17069/2020\|EPI_ISL_750741\|202 | ···················· | ···················· | ····················· | ···················· | ···················· | ···················· |
| AA | 1379 | hCoV-19/Denmark/DCGC-17071/2020\|EPI_ISL_750742\|202 | ···················· | ···················· | ····················· | ···················· | ···················· | ···················· |
| AA | 1380 | hCoV-19/Denmark/DCGC-17073/2020\|EPI_ISL_750743\|202 | ···················· | ···················· | ····················· | ···················· | ···················· | ···················· |
| AA | 1381 | hCoV-19/Denmark/DCGC-17076/2020\|EPI_ISL_750744\|202 | ···················· | ···················· | ····················· | ···················· | ···················· | ···················· |
| AA | 1382 | hCoV-19/Denmark/DCGC-17079/2020\|EPI_ISL_750745\|202 | ···················· | ···················· | ····················· | ···················· | ···················· | ···················· |
| AA | 1383 | hCoV-19/Denmark/DCGC-17080/2020\|EPI_ISL_750746\|202 | ···················· | ···················· | ····················· | ···················· | ···················· | ···················· |
| AA | 1384 | hCoV-19/Denmark/DCGC-17082/2020\|EPI_ISL_750747\|202 | ···················· | ···················· | ····················· | ···················· | ···················· | ···················· |
| AA | 1385 | hCoV-19/Denmark/DCGC-17083/2020\|EPI_ISL_750748\|202 | ···················· | ···················· | ····················· | ···················· | ···················· | ···················· |
| AA | 1386 | hCoV-19/Denmark/DCGC-17087/2020\|EPI_ISL_750749\|202 | ···················· | ···················· | ····················· | ···················· | ···················· | ···················· |
| AA | 1387 | hCoV-19/Denmark/DCGC-17088/2020\|EPI_ISL_750750\|202 | ···················· | ···················· | ····················· | ···················· | ···················· | ···················· |
| AA | 1388 | hCoV-19/Denmark/DCGC-17090/2020\|EPI_ISL_750751\|202 | ···················· | ···················· | ····················· | ···················· | ···················· | ···················· |
| AA | 1389 | hCoV-19/Denmark/DCGC-17091/2020\|EPI_ISL_750752\|202 | ···················· | ···················· | ····················· | ···················· | ···················· | ···················· |
| AA | 1390 | hCoV-19/Denmark/DCGC-17094/2020\|EPI_ISL_750753\|202 | ···················· | ···················· | ····················· | ···················· | ···················· | ···················· |
| AA | 1391 | hCoV-19/Denmark/DCGC-17095/2020\|EPI_ISL_750754\|202 | ···················· | ···················· | ····················· | ···················· | ···················· | ···················· |
| AA | 1392 | hCoV-19/Denmark/DCGC-17097/2020\|EPI_ISL_750755\|202 | ···················· | ···················· | ····················· | ···················· | ···················· | ···················· |
| AA | 1393 | hCoV-19/Denmark/DCGC-17098/2020\|EPI_ISL_750756\|202 | ···················· | ···················· | ····················· | ···················· | ···················· | ···················· |
| AA | 1394 | hCoV-19/Denmark/DCGC-17099/2020\|EPI_ISL_750757\|202 | ···················· | ···················· | ····················· | ···················· | ···················· | ···················· |
| AA | 1395 | hCoV-19/Denmark/DCGC-17100/2020\|EPI_ISL_750758\|202 | ···················· | ···················· | ····················· | ···················· | ···················· | ···················· |
| AA | 1396 | hCoV-19/Denmark/DCGC-17101/2020\|EPI_ISL_750759\|202 | ···················· | ···················· | ····················· | ···················· | ···················· | ···················· |
| AA | 1397 | hCoV-19/Denmark/DCGC-17105/2020\|EPI_ISL_750760\|202 | ···················· | ···················· | ····················· | ···················· | ···················· | ···················· |
| AA | 1398 | hCoV-19/Denmark/DCGC-17107/2020\|EPI_ISL_750761\|202 | ···················· | ···················· | ····················· | ···················· | ···················· | ···················· |
| AA | 1399 | hCoV-19/Denmark/DCGC-17108/2020\|EPI_ISL_750762\|202 | ···················· | ···················· | ····················· | ···················· | ···················· | ···················· |
| AA | 1400 | hCoV-19/Denmark/DCGC-17112/2020\|EPI_ISL_750763\|202 | ···················· | ···················· | ····················· | ···················· | ···················· | ···················· |
| AA | 1401 | hCoV-19/Denmark/DCGC-17114/2020\|EPI_ISL_750764\|202 | ···················· | ···················· | ····················· | ···················· | ···················· | ···················· |
| AA | 1402 | hCoV-19/Denmark/DCGC-17117/2020\|EPI_ISL_750765\|202 | ···················· | ···················· | ····················· | ···················· | ···················· | ···················· |
| AA | 1403 | hCoV-19/Denmark/DCGC-17118/2020\|EPI_ISL_750766\|202 | ···················· | ···················· | ····················· | ···················· | ···················· | ···················· |
| AA | 1404 | hCoV-19/Denmark/DCGC-17119/2020\|EPI_ISL_750767\|202 | ···················· | ···················· | ····················· | ···················· | ···················· | ···················· |
| AA | 1405 | hCoV-19/Denmark/DCGC-17120/2020\|EPI_ISL_750768\|202 | ···················· | ···················· | ····················· | ···················· | ···················· | ···················· |
| AA | 1406 | hCoV-19/Denmark/DCGC-17121/2020\|EPI_ISL_750769\|202 | ···················· | ···················· | ····················· | ···················· | ···················· | ···················· |
| AA | 1407 | hCoV-19/Denmark/DCGC-17122/2020\|EPI_ISL_750770\|202 | ···················· | ···················· | ····················· | ···················· | ···················· | ···················· |
| AA | 1408 | hCoV-19/Denmark/DCGC-17124/2020\|EPI_ISL_750771\|202 | ···················· | ·A·················· | ····················· | ···················· | ···················· | ···················· |
| AA | 1409 | hCoV-19/Denmark/DCGC-17126/2020\|EPI_ISL_750772\|202 | ···················· | ···················· | ····················· | ···················· | ···················· | ···················· |
| AA | 1410 | hCoV-19/Denmark/DCGC-17128/2020\|EPI_ISL_750773\|202 | ···················· | ···················· | ····················· | ···················· | ···················· | ···················· |
| AA | 1411 | hCoV-19/Denmark/DCGC-17132/2020\|EPI_ISL_750774\|202 | ···················· | ···················· | ····················· | ···················· | ···················· | ···················· |
| AA | 1412 | hCoV-19/Denmark/DCGC-17133/2020\|EPI_ISL_750775\|202 | ···················· | ···················· | ····················· | ···················· | ···················· | ···················· |
| AA | 1413 | hCoV-19/Denmark/DCGC-17134/2020\|EPI_ISL_750776\|202 | ···················· | ···················· | ····················· | ···················· | ···················· | ···················· |
| AA | 1414 | hCoV-19/Denmark/DCGC-17135/2020\|EPI_ISL_750777\|202 | ···················· | ···················· | ····················· | ···················· | ···················· | ···················· |
| AA | 1415 | hCoV-19/Denmark/DCGC-17136/2020\|EPI_ISL_750778\|202 | ···················· | ···················· | ····················· | ···················· | ···················· | ···················· |
| AA | 1416 | hCoV-19/Denmark/DCGC-17137/2020\|EPI_ISL_750779\|202 | ···················· | ···················· | ····················· | ···················· | ···················· | ···················· |
| AA | 1417 | hCoV-19/Denmark/DCGC-17138/2020\|EPI_ISL_750780\|202 | ···················· | ···················· | ····················· | ···················· | ···················· | ···················· |
| AA | 1418 | hCoV-19/Denmark/DCGC-17139/2020\|EPI_ISL_750781\|202 | ···················· | ···················· | ····················· | ···················· | ···················· | ···················· |
| AA | 1419 | hCoV-19/Denmark/DCGC-17140/2020\|EPI_ISL_750782\|202 | ···················· | ···················· | ····················· | ···················· | ···················· | ···················· |
| AA | 1420 | hCoV-19/Denmark/DCGC-17141/2020\|EPI_ISL_750783\|202 | ···················· | ···················· | ····················· | ···················· | ···················· | ···················· |
| AA | 1421 | hCoV-19/Denmark/DCGC-17143/2020\|EPI_ISL_750784\|202 | ···················· | ···················· | ····················· | ···················· | ···················· | ···················· |
| AA | 1422 | hCoV-19/Denmark/DCGC-17144/2020\|EPI_ISL_750785\|202 | ···················· | ···················· | ····················· | ···················· | ···················· | ···················· |
| AA | 1423 | hCoV-19/Denmark/DCGC-17147/2020\|EPI_ISL_750786\|202 | ···················· | ···················· | ····················· | ···················· | ···················· | ···················· |
| AA | 1424 | hCoV-19/Denmark/DCGC-17148/2020\|EPI_ISL_750787\|202 | ···················· | ···················· | ····················· | ···················· | ···················· | ···················· |
| AA | 1425 | hCoV-19/Denmark/DCGC-17150/2020\|EPI_ISL_750788\|202 | ···················· | ···················· | ····················· | ···················· | ···················· | ···················· |
| AA | 1426 | hCoV-19/Denmark/DCGC-17151/2020\|EPI_ISL_750789\|202 | ···················· | ···················· | ····················· | ···················· | ···················· | ···················· |
| AA | 1427 | hCoV-19/Denmark/DCGC-17152/2020\|EPI_ISL_750790\|202 | ···················· | ···················· | ····················· | ···················· | ···················· | ···················· |
| AA | 1428 | hCoV-19/Denmark/DCGC-17153/2020\|EPI_ISL_750791\|202 | ···················· | ···················· | ····················· | ···················· | ···················· | ···················· |
| AA | 1429 | hCoV-19/Denmark/DCGC-17155/2020\|EPI_ISL_750792\|202 | ···················· | ···················· | ····················· | ···················· | ···················· | ···················· |
| AA | 1430 | hCoV-19/Denmark/DCGC-17156/2020\|EPI_ISL_750793\|202 | ···················· | ···················· | ····················· | ···················· | ···················· | ···················· |
| AA | 1431 | hCoV-19/Denmark/DCGC-17158/2020\|EPI_ISL_750794\|202 | ···················· | ···················· | ····················· | ···················· | ···················· | ···················· |
| AA | 1432 | hCoV-19/Denmark/DCGC-17159/2020\|EPI_ISL_750795\|202 | ···················· | ···················· | ····················· | ···················· | ···················· | ···················· |
| AA | 1433 | hCoV-19/Denmark/DCGC-17164/2020\|EPI_ISL_750796\|202 | ···················· | ···················· | ····················· | ···················· | ···················· | ···················· |
| AA | 1434 | hCoV-19/Denmark/DCGC-17165/2020\|EPI_ISL_750797\|202 | ···················· | ···················· | ····················· | ···················· | ···················· | ···················· |
| AA | 1435 | hCoV-19/Denmark/DCGC-17168/2020\|EPI_ISL_750798\|202 | ···················· | ···················· | ····················· | ···················· | ···················· | ···················· |
| AA | 1436 | hCoV-19/Denmark/DCGC-17169/2020\|EPI_ISL_750799\|202 | ···················· | ···················· | ····················· | ···················· | ···················· | ···················· |
| AA | 1437 | hCoV-19/England/204941496/2020\|EPI_ISL_754300\|2020 | ···················· | ·········M·········· | ····················· | ···················· | ···················· | ···················· |
| AA | 1438 | hCoV-19/England/204941554/2020\|EPI_ISL_754301\|2020 | ···················· | ···················· | ····················· | ···················· | ···················· | ···················· |
| AA | 1439 | hCoV-19/England/204941557/2020\|EPI_ISL_754302\|2020 | ···················· | ···················· | ····················· | ···················· | ···················· | ···················· |
| AA | 1440 | hCoV-19/England/204941558/2020\|EPI_ISL_754303\|2020 | ···················· | ···················· | ····················· | ···················· | ···················· | ···················· |
| AA | 1441 | hCoV-19/England/204941562/2020\|EPI_ISL_754304\|2020 | ···················· | ···················· | ····················· | ···················· | ···················· | ···················· |
| AA | 1442 | hCoV-19/USA/MA-CDC-STM-P006/2020\|EPI_ISL_755600\|20 | ···················· | ···················· | ····················· | ···················· | ···················· | ···················· |

| | | | | | | | | |
|---|---|---|---|---|---|---|---|---|
| AA | 1443 | hCoV-19/USA/FL-CDC-STM-P007/2020\|EPI_ISL_755601\|20 | ···················· | ···················· | ···················· | ···················· | ···················· | ···················· |
| AA | 1444 | hCoV-19/USA/CA-CDC-STM-P008/2020\|EPI_ISL_755602\|20 | ···················· | ···················· | ···················· | ···················· | ···················· | ···················· |
| AA | 1445 | hCoV-19/USA/MA-CDC-STM-P009/2020\|EPI_ISL_755603\|20 | ···················· | ···················· | ···················· | ···················· | ···················· | ···················· |
| AA | 1446 | hCoV-19/USA/FL-CDC-STM-P011/2020\|EPI_ISL_755604\|20 | ···················· | ···················· | ···················· | ···················· | ···················· | ···················· |
| AA | 1447 | hCoV-19/USA/FL-CDC-STM-P013/2020\|EPI_ISL_755605\|20 | ···················· | ···················· | ···················· | ···················· | ···················· | ···················· |
| AA | 1448 | hCoV-19/USA/FL-CDC-STM-P014/2020\|EPI_ISL_755606\|20 | ···················· | ···················· | ···················· | ···················· | ···················· | ···················· |
| AA | 1449 | hCoV-19/USA/FL-CDC-STM-P015/2020\|EPI_ISL_755607\|20 | ···················· | ···················· | ···················· | ···················· | ···················· | ···················· |
| AA | 1450 | hCoV-19/USA/MA-CDC-STM-P018/2020\|EPI_ISL_755609\|20 | ···················· | ···················· | ···················· | ···················· | ···················· | ···················· |
| AA | 1451 | hCoV-19/USA/PA-CDC-STM-P020/2020\|EPI_ISL_755610\|20 | ···················· | ···················· | ···················· | ···················· | ···················· | ···················· |
| AA | 1452 | hCoV-19/USA/MA-CDC-STM-P021/2020\|EPI_ISL_755611\|20 | ···················· | ···················· | ···················· | ···················· | ···················· | ···················· |
| AA | 1453 | hCoV-19/USA/IN-CDC-STM-P053/2020\|EPI_ISL_755616\|20 | ···················· | ···················· | ···················· | ···················· | ···················· | ···················· |
| AA | 1454 | hCoV-19/USA/MA-CDC-STM-P056/2020\|EPI_ISL_755617\|20 | ···················· | ···················· | ···················· | ···················· | ···················· | ···················· |
| AA | 1455 | hCoV-19/New Zealand/20CV0675/2020\|EPI_ISL_755622\|2 | ···················· | ···················· | ···················· | ···················· | ···················· | ···················· |
| AA | 1456 | hCoV-19/New Zealand/20CV0697/2020\|EPI_ISL_755630\|2 | ···················· | ···················· | ···················· | ···················· | ···················· | ···················· |
| AA | 1457 | hCoV-19/New Zealand/20CV0701/2020\|EPI_ISL_755631\|2 | ···················· | ···················· | ···················· | ···················· | ···················· | ···················· |
| AA | 1458 | hCoV-19/New Zealand/20CV0704/2020\|EPI_ISL_755633\|2 | ···················· | ·A·················· | ···················· | ···················· | ···················· | ···A················ |
| AA | 1459 | hCoV-19/New Zealand/20CV0709/2020\|EPI_ISL_755634\|2 | ···················· | ·A·················· | ···················· | ···················· | ···················· | ···A················ |
| AA | 1460 | hCoV-19/New Zealand/20CV0715/2020\|EPI_ISL_755635\|2 | ···················· | ···················· | ···················· | ···················· | ···················· | ···················· |
| AA | 1461 | hCoV-19/England/QEUH-C7C2B2/2020\|EPI_ISL_756923\|20 | ···················· | ···················· | ···················· | ···················· | ···················· | ···················· |
| AA | 1462 | hCoV-19/England/MILK-C7B8C0/2020\|EPI_ISL_756924\|20 | ···················· | ···················· | ···················· | ···················· | ······T············· | ···················· |
| AA | 1463 | hCoV-19/England/MILK-C7B74B/2020\|EPI_ISL_756925\|20 | ···················· | ···················· | ···················· | ···················· | ···················· | ···················· |
| AA | 1464 | hCoV-19/England/QEUH-C7C22B/2020\|EPI_ISL_756940\|20 | ···················· | ···················· | ···················· | ···················· | ···················· | ···················· |
| AA | 1465 | hCoV-19/England/MILK-C7B20E/2020\|EPI_ISL_756944\|20 | ···················· | ···················· | ···················· | ···················· | ···················· | ···················· |
| AA | 1466 | hCoV-19/England/QEUH-C7FFC7/2020\|EPI_ISL_756947\|20 | ···················· | ···················· | ···················· | ···················· | ···················· | ···················· |
| AA | 1467 | hCoV-19/England/QEUH-C7D873/2020\|EPI_ISL_756951\|20 | ···················· | ···················· | ···················· | ···················· | ···················· | ···················· |
| AA | 1468 | hCoV-19/England/QEUH-C80109/2020\|EPI_ISL_756953\|20 | ···················· | ···················· | ···················· | ···················· | ···················· | ···················· |
| AA | 1469 | hCoV-19/England/MILK-C7AA25/2020\|EPI_ISL_756956\|20 | ···················· | ·A·················· | ···················· | ···················· | ···················· | ···A················ |
| AA | 1470 | hCoV-19/England/MILK-C78289/2020\|EPI_ISL_756958\|20 | ···················· | ···················· | ···················· | ···················· | ···················· | ···················· |
| AA | 1471 | hCoV-19/England/QEUH-C7C2A3/2020\|EPI_ISL_756959\|20 | ···················· | ···················· | ···················· | ···················· | ···················· | ···················· |
| AA | 1472 | hCoV-19/England/MILK-C7B866/2020\|EPI_ISL_756963\|20 | ·······T············ | ·A·················· | ···················· | ···················· | ···················· | ···················· |
| AA | 1473 | hCoV-19/Denmark/DCGC-20359/2020\|EPI_ISL_759104\|202 | ···················· | ·A·················· | ···················· | ···················· | ···················· | ···A················ |
| AA | 1474 | hCoV-19/Denmark/DCGC-20361/2020\|EPI_ISL_759105\|202 | ···················· | ···················· | ···················· | ···················· | ···················· | ···················· |
| AA | 1475 | hCoV-19/Denmark/DCGC-20365/2020\|EPI_ISL_759106\|202 | ···················· | ···················· | ···················· | ···················· | ···················· | ···················· |
| AA | 1476 | hCoV-19/Denmark/DCGC-20366/2020\|EPI_ISL_759107\|202 | ···················· | ···················· | ···················· | ···················· | ···················· | ···················· |
| AA | 1477 | hCoV-19/Denmark/DCGC-20367/2020\|EPI_ISL_759108\|202 | ···················· | ···················· | ···················· | ···················· | ···················· | ···················· |
| AA | 1478 | hCoV-19/Denmark/DCGC-20368/2020\|EPI_ISL_759109\|202 | ···················· | ···················· | ···················· | ···················· | ···················· | ···················· |
| AA | 1479 | hCoV-19/Denmark/DCGC-20369/2020\|EPI_ISL_759110\|202 | ···················· | ···················· | ···················· | ···················· | ···················· | ···················· |
| AA | 1480 | hCoV-19/Denmark/DCGC-20372/2020\|EPI_ISL_759111\|202 | ···················· | ···················· | ···················· | ···················· | ···················· | ···················· |
| AA | 1481 | hCoV-19/Denmark/DCGC-20374/2020\|EPI_ISL_759112\|202 | ···················· | ···················· | ···················· | ···················· | ···················· | ···················· |
| AA | 1482 | hCoV-19/Denmark/DCGC-20375/2020\|EPI_ISL_759113\|202 | ···················· | ···················· | ···················· | ···················· | ···················· | ···················· |
| AA | 1483 | hCoV-19/Denmark/DCGC-20376/2020\|EPI_ISL_759114\|202 | ···················· | ···················· | ···················· | ···················· | ···················· | ···················· |
| AA | 1484 | hCoV-19/Denmark/DCGC-20377/2020\|EPI_ISL_759115\|202 | ···················· | ···················· | ···················· | ···················· | ···················· | ···················· |
| AA | 1485 | hCoV-19/Denmark/DCGC-20379/2020\|EPI_ISL_759116\|202 | ···················· | ···················· | ···················· | ···················· | ···················· | ···················· |
| AA | 1486 | hCoV-19/Denmark/DCGC-20382/2020\|EPI_ISL_759117\|202 | ···················· | ···················· | ···················· | ···················· | ···················· | ···················· |
| AA | 1487 | hCoV-19/Denmark/DCGC-20386/2020\|EPI_ISL_759118\|202 | ···················· | ···················· | ···················· | ···················· | ···················· | ···················· |
| AA | 1488 | hCoV-19/Denmark/DCGC-20390/2020\|EPI_ISL_759119\|202 | ···················· | ···················· | ···················· | ···················· | ···················· | ···················· |
| AA | 1489 | hCoV-19/Denmark/DCGC-20392/2020\|EPI_ISL_759120\|202 | ···················· | ···················· | ···················· | ···················· | ···················· | ···················· |
| AA | 1490 | hCoV-19/Denmark/DCGC-20393/2020\|EPI_ISL_759121\|202 | ···················· | ···················· | ···················· | ···················· | ···················· | ···················· |
| AA | 1491 | hCoV-19/Denmark/DCGC-20394/2020\|EPI_ISL_759122\|202 | ···················· | ···················· | ···················· | ···················· | ···················· | ···················· |
| AA | 1492 | hCoV-19/Denmark/DCGC-20395/2020\|EPI_ISL_759123\|202 | ···················· | ···················· | ···················· | ···················· | ···················· | ···················· |
| AA | 1493 | hCoV-19/Denmark/DCGC-20397/2020\|EPI_ISL_759124\|202 | ···················· | ···················· | ···················· | ···················· | ···················· | ···················· |
| AA | 1494 | hCoV-19/Denmark/DCGC-20402/2020\|EPI_ISL_759125\|202 | ···················· | ···················· | ···················· | ···················· | ···················· | ···················· |
| AA | 1495 | hCoV-19/Denmark/DCGC-20403/2020\|EPI_ISL_759126\|202 | ···················· | ···················· | ···················· | ···················· | ···················· | ···················· |
| AA | 1496 | hCoV-19/Denmark/DCGC-20404/2020\|EPI_ISL_759127\|202 | ···················· | ···················· | ···················· | ···················· | ···················· | ···················· |
| AA | 1497 | hCoV-19/Denmark/DCGC-20406/2020\|EPI_ISL_759128\|202 | ···················· | ···················· | ···················· | ···················· | ···················· | ···················· |
| AA | 1498 | hCoV-19/Denmark/DCGC-20407/2020\|EPI_ISL_759129\|202 | ···················· | ···················· | ···················· | ···················· | ···················· | ···················· |
| AA | 1499 | hCoV-19/Denmark/DCGC-20408/2020\|EPI_ISL_759130\|202 | ···················· | ···················· | ···················· | ···················· | ···················· | ···················· |
| AA | 1500 | hCoV-19/Denmark/DCGC-20410/2020\|EPI_ISL_759131\|202 | ···················· | ···················· | ···················· | ···················· | ···················· | ···················· |
| AA | 1501 | hCoV-19/Denmark/DCGC-20411/2020\|EPI_ISL_759132\|202 | ···················· | ···················· | ···················· | ···················· | ···················· | ···················· |
| AA | 1502 | hCoV-19/Denmark/DCGC-20414/2020\|EPI_ISL_759133\|202 | ···················· | ···················· | ···················· | ···················· | ···················· | ···················· |
| AA | 1503 | hCoV-19/Denmark/DCGC-20415/2020\|EPI_ISL_759134\|202 | ···················· | ···················· | ···················· | ···················· | ···················· | ···················· |
| AA | 1504 | hCoV-19/Denmark/DCGC-20417/2020\|EPI_ISL_759135\|202 | ···················· | ·A·················· | ···················· | ···················· | ···················· | ···A················ |
| AA | 1505 | hCoV-19/Denmark/DCGC-20418/2020\|EPI_ISL_759136\|202 | ···················· | ···················· | ···················· | ···················· | ···················· | ···················· |
| AA | 1506 | hCoV-19/Denmark/DCGC-20419/2020\|EPI_ISL_759137\|202 | ···················· | ···················· | ···················· | ···················· | ···················· | ···················· |
| AA | 1507 | hCoV-19/Denmark/DCGC-20422/2020\|EPI_ISL_759138\|202 | ···················· | ···················· | ···················· | ···················· | ···················· | ···················· |
| AA | 1508 | hCoV-19/Denmark/DCGC-20424/2020\|EPI_ISL_759139\|202 | ···················· | ···················· | ···················· | ···················· | ···················· | ···················· |
| AA | 1509 | hCoV-19/Denmark/DCGC-20425/2020\|EPI_ISL_759140\|202 | ···················· | ···················· | ···················· | ···················· | ···················· | ···················· |
| AA | 1510 | hCoV-19/Denmark/DCGC-20428/2020\|EPI_ISL_759141\|202 | ···················· | ···················· | ···················· | ···················· | ···················· | ···················· |
| AA | 1511 | hCoV-19/Denmark/DCGC-20430/2020\|EPI_ISL_759142\|202 | ···················· | ···················· | ···················· | ···················· | ···················· | ···················· |
| AA | 1512 | hCoV-19/Denmark/DCGC-20432/2020\|EPI_ISL_759143\|202 | ···················· | ···················· | ···················· | ···················· | ···················· | ···················· |
| AA | 1513 | hCoV-19/Denmark/DCGC-20434/2020\|EPI_ISL_759144\|202 | ···················· | ·A·················· | ···················· | ···················· | ···················· | ···A················ |
| AA | 1514 | hCoV-19/Denmark/DCGC-20435/2020\|EPI_ISL_759145\|202 | ···················· | ···················· | ···················· | ···················· | ···················· | ···················· |
| AA | 1515 | hCoV-19/Denmark/DCGC-20436/2020\|EPI_ISL_759146\|202 | ···················· | ·A·················· | ···················· | ···················· | ···················· | ···A················ |
| AA | 1516 | hCoV-19/Denmark/DCGC-20437/2020\|EPI_ISL_759147\|202 | ···················· | ·A·················· | ···················· | ···················· | ···················· | ···A················ |
| AA | 1517 | hCoV-19/Denmark/DCGC-20438/2020\|EPI_ISL_759148\|202 | ···················· | ·A·················· | ···················· | ···················· | ···················· | ···A················ |
| AA | 1518 | hCoV-19/Denmark/DCGC-20440/2020\|EPI_ISL_759149\|202 | ···················· | ···················· | ···················· | ···················· | ···················· | ···················· |
| AA | 1519 | hCoV-19/Denmark/DCGC-20441/2020\|EPI_ISL_759150\|202 | ···················· | ···················· | ···················· | ···················· | ···················· | ···················· |
| AA | 1520 | hCoV-19/Denmark/DCGC-20443/2020\|EPI_ISL_759151\|202 | ···················· | ···················· | ···················· | ···················· | ···················· | ···················· |
| AA | 1521 | hCoV-19/Denmark/DCGC-20445/2020\|EPI_ISL_759152\|202 | ···················· | ···················· | ···················· | ···················· | ···················· | ···················· |
| AA | 1522 | hCoV-19/Denmark/DCGC-20446/2020\|EPI_ISL_759153\|202 | ···················· | ···················· | ···················· | ···················· | ···················· | ···················· |

| | | | | | | | |
|---|---|---|---|---|---|---|---|
| AA | 1523 hCoV-19/Denmark/DCGC-20447/2020\|EPI_ISL_759154\|202 | ···················· | ···················· | ···················· | ···················· | ···················· | ···················· |
| AA | 1524 hCoV-19/Denmark/DCGC-20448/2020\|EPI_ISL_759155\|202 | ···················· | ···················· | ···················· | ···················· | ···················· | ···················· |
| AA | 1525 hCoV-19/Denmark/DCGC-20449/2020\|EPI_ISL_759156\|202 | ···················· | ···················· | ···················· | ···················· | ···················· | ···················· |
| AA | 1526 hCoV-19/Denmark/DCGC-20450/2020\|EPI_ISL_759157\|202 | ···················· | ···················· | ···················· | ···················· | ···················· | ···················· |
| AA | 1527 hCoV-19/Denmark/DCGC-20451/2020\|EPI_ISL_759158\|202 | ···················· | ···················· | ···················· | ···················· | ···················· | ···················· |
| AA | 1528 hCoV-19/Denmark/DCGC-20452/2020\|EPI_ISL_759159\|202 | ···················· | ···················· | ···················· | ···················· | ···················· | ···················· |
| AA | 1529 hCoV-19/Denmark/DCGC-20453/2020\|EPI_ISL_759160\|202 | ···················· | ···················· | ···················· | ···················· | ···················· | ···················· |
| AA | 1530 hCoV-19/Denmark/DCGC-20455/2020\|EPI_ISL_759161\|202 | ···················· | ···················· | ···················· | ···················· | ···················· | ···················· |
| AA | 1531 hCoV-19/Denmark/DCGC-20456/2020\|EPI_ISL_759162\|202 | ···················· | ···················· | ···················· | ···················· | ···················· | ···················· |
| AA | 1532 hCoV-19/Denmark/DCGC-20457/2020\|EPI_ISL_759163\|202 | ···················· | ···················· | ···················· | ···················· | ···················· | ···················· |
| AA | 1533 hCoV-19/Denmark/DCGC-20460/2020\|EPI_ISL_759164\|202 | ···················· | ···················· | ···················· | ···················· | ···················· | ···················· |
| AA | 1534 hCoV-19/Denmark/DCGC-20461/2020\|EPI_ISL_759165\|202 | ···················· | ···················· | ···················· | ···················· | ···················· | ···················· |
| AA | 1535 hCoV-19/Denmark/DCGC-20465/2020\|EPI_ISL_759168\|202 | ···················· | ···················· | ···················· | ···················· | ···················· | ···················· |
| AA | 1536 hCoV-19/Denmark/DCGC-20467/2020\|EPI_ISL_759169\|202 | ···················· | ···················· | ···················· | ···················· | ···················· | ···················· |
| AA | 1537 hCoV-19/Denmark/DCGC-19110/2020\|EPI_ISL_748507\|202 | ···················· | ···················· | ···················· | ···················· | ···················· | ···················· |
| AA | 1538 hCoV-19/Denmark/DCGC-18235/2020\|EPI_ISL_749831\|202 | ···················· | ···················· | ···················· | ···················· | ···················· | ···················· |
| AA | 1539 hCoV-19/Denmark/DCGC-18236/2020\|EPI_ISL_749832\|202 | ···················· | ···················· | ···················· | ···················· | ···················· | ···················· |
| AA | 1540 hCoV-19/Denmark/DCGC-18237/2020\|EPI_ISL_749833\|202 | ···················· | ···················· | ···················· | ···················· | ···················· | ···················· |
| AA | 1541 hCoV-19/Denmark/DCGC-18240/2020\|EPI_ISL_749835\|202 | ···················· | ···················· | ···················· | ···················· | ···················· | ···················· |
| AA | 1542 hCoV-19/Denmark/DCGC-18241/2020\|EPI_ISL_749836\|202 | ···················· | ···················· | ···················· | ···················· | ···················· | ···················· |
| AA | 1543 hCoV-19/Denmark/DCGC-18242/2020\|EPI_ISL_749837\|202 | ···················· | ···················· | ···················· | ···················· | ···················· | ···················· |
| AA | 1544 hCoV-19/Denmark/DCGC-18244/2020\|EPI_ISL_749838\|202 | ···················· | ···················· | ···················· | ···················· | ···················· | ···················· |
| AA | 1545 hCoV-19/Uruguay/TYT-M135/2020\|EPI_ISL_750820\|2020- | ···················· | ···················· | ···················· | ···················· | ···················· | ···················· |
| AA | 1546 hCoV-19/Denmark/DCGC-17201/2020\|EPI_ISL_750821\|202 | ···················· | ···················· | ···················· | ···················· | ···················· | ···················· |
| AA | 1547 hCoV-19/Denmark/DCGC-17202/2020\|EPI_ISL_750822\|202 | ···················· | ···················· | ···················· | ···················· | ···················· | ···················· |
| AA | 1548 hCoV-19/Denmark/DCGC-17205/2020\|EPI_ISL_750823\|202 | ···················· | ···················· | ···················· | ···················· | ···················· | ···················· |
| AA | 1549 hCoV-19/Denmark/DCGC-17206/2020\|EPI_ISL_750824\|202 | ···················· | ···················· | ···················· | ···················· | ···················· | ···················· |
| AA | 1550 hCoV-19/Denmark/DCGC-17207/2020\|EPI_ISL_750825\|202 | ···················· | ···················· | ···················· | ···················· | ···················· | ···················· |
| AA | 1551 hCoV-19/USA/HI-H200457/2020\|EPI_ISL_753000\|2020-09 | ···················· | ·A·················· | ···················· | ···················· | ···················· | ···················· |
| AA | 1552 hCoV-19/USA/HI-H200459/2020\|EPI_ISL_753002\|2020-10 | ···················· | ·A·················· | ···················· | ···················· | ···················· | ···················· |
| AA | 1553 hCoV-19/USA/HI-H200460/2020\|EPI_ISL_753003\|2020-10 | ···················· | ·A·················· | ···················· | ···················· | ···················· | ···················· |
| AA | 1554 hCoV-19/USA/HI-H200461/2020\|EPI_ISL_753004\|2020-10 | ···················· | ·A·················· | ···················· | ···················· | ···················· | ···················· |
| AA | 1555 hCoV-19/USA/HI-H200462/2020\|EPI_ISL_753005\|2020-10 | ···················· | ·A·················· | ···················· | ···················· | ···················· | ···················· |
| AA | 1556 hCoV-19/USA/HI-H200463/2020\|EPI_ISL_753006\|2020-10 | ···················· | ·A·················· | ···················· | ···················· | ···················· | ···················· |
| AA | 1557 hCoV-19/USA/HI-H200464/2020\|EPI_ISL_753007\|2020-10 | ···················· | ·A·················· | ···················· | ···················· | ···················· | ···················· |
| AA | 1558 hCoV-19/USA/HI-H200465/2020\|EPI_ISL_753008\|2020-10 | ···················· | ·A·················· | ···················· | ···················· | ···················· | ···················· |
| AA | 1559 hCoV-19/USA/HI-H200466/2020\|EPI_ISL_753009\|2020-10 | ···················· | ·A·················· | ···················· | ···················· | ···················· | ···················· |
| AA | 1560 hCoV-19/USA/HI-H200467/2020\|EPI_ISL_753010\|2020-10 | ···················· | ·A·················· | ···················· | ···················· | ···················· | ···················· |
| AA | 1561 hCoV-19/USA/HI-H200468/2020\|EPI_ISL_753011\|2020-10 | ···················· | ·A·················· | ···················· | ···················· | ···················· | ···················· |
| AA | 1562 hCoV-19/USA/HI-H200469/2020\|EPI_ISL_753012\|2020-10 | ···················· | ·A·················· | ···················· | ···················· | ···················· | ···················· |
| AA | 1563 hCoV-19/USA/HI-H200470/2020\|EPI_ISL_753013\|2020-10 | ···················· | ·A·················· | ···················· | ···················· | ···················· | ···················· |
| AA | 1564 hCoV-19/USA/HI-H200471/2020\|EPI_ISL_753014\|2020-10 | ···················· | ·A·················· | ···················· | ···················· | ···················· | ···················· |
| AA | 1565 hCoV-19/USA/HI-H200472/2020\|EPI_ISL_753015\|2020-10 | ···················· | ·A·················· | ···················· | ···················· | ···················· | ···················· |
| AA | 1566 hCoV-19/USA/HI-H200473/2020\|EPI_ISL_753016\|2020-10 | ···················· | ·A·················· | ···················· | ···················· | ···················· | ···················· |
| AA | 1567 hCoV-19/USA/HI-H200474/2020\|EPI_ISL_753017\|2020-10 | ···················· | ·A·················· | ···················· | ···················· | ···················· | ···················· |
| AA | 1568 hCoV-19/USA/HI-H200475/2020\|EPI_ISL_753018\|2020-10 | ···················· | ·A·················· | ···················· | ···················· | ···················· | ···················· |
| AA | 1569 hCoV-19/USA/HI-H200476/2020\|EPI_ISL_753019\|2020-10 | ···················· | ···················· | ···················· | ···················· | ···················· | ···················· |
| AA | 1570 hCoV-19/USA/HI-H200477/2020\|EPI_ISL_753020\|2020-08 | ···················· | ·A·················· | ···················· | ···················· | ···················· | ···················· |
| AA | 1571 hCoV-19/USA/HI-H200479/2020\|EPI_ISL_753021\|2020-08 | ···················· | ·A·················· | ···················· | ···················· | ···················· | ···················· |
| AA | 1572 hCoV-19/USA/HI-H200480/2020\|EPI_ISL_753022\|2020-09 | ···················· | ·A·················· | ···················· | ···················· | ···················· | ···················· |
| AA | 1573 hCoV-19/USA/HI-H200481/2020\|EPI_ISL_753023\|2020-09 | ···················· | ·A·················· | ···················· | ···················· | ···················· | ···················· |
| AA | 1574 hCoV-19/USA/HI-H200482/2020\|EPI_ISL_753024\|2020-09 | ···················· | ·A·················· | ···················· | ···················· | ···················· | ···················· |
| AA | 1575 hCoV-19/USA/HI-H200483/2020\|EPI_ISL_753025\|2020-09 | ···················· | ·A·················· | ···················· | ···················· | ···················· | ···················· |
| AA | 1576 hCoV-19/USA/HI-H200485/2020\|EPI_ISL_753026\|2020-09 | ···················· | ·A·················· | ···················· | ···················· | ···················· | ···················· |
| AA | 1577 hCoV-19/USA/HI-H200486/2020\|EPI_ISL_753027\|2020-09 | ···················· | ·A·················· | ···················· | ···················· | ···················· | ···················· |
| AA | 1578 hCoV-19/USA/HI-H200487/2020\|EPI_ISL_753028\|2020-09 | ···················· | ·A·················· | ···················· | ···················· | ···················· | ···················· |
| AA | 1579 hCoV-19/USA/HI-H200488/2020\|EPI_ISL_753029\|2020-09 | ···················· | ·A·················· | ···················· | ···················· | ···················· | ···················· |
| AA | 1580 hCoV-19/USA/HI-H200490/2020\|EPI_ISL_753030\|2020-09 | ···················· | ·A·················· | ···················· | ···················· | ···················· | ···················· |
| AA | 1581 hCoV-19/USA/HI-H200492/2020\|EPI_ISL_753031\|2020-10 | ···················· | ·A·················· | ···················· | ···················· | ···················· | ···················· |
| AA | 1582 hCoV-19/USA/HI-H200493/2020\|EPI_ISL_753032\|2020-10 | ···················· | ·A·················· | ···················· | ···················· | ···················· | ···················· |
| AA | 1583 hCoV-19/USA/HI-H200494/2020\|EPI_ISL_753033\|2020-10 | ···················· | ·A·················· | ···················· | ···················· | ···················· | ···················· |
| AA | 1584 hCoV-19/USA/HI-H200495/2020\|EPI_ISL_753034\|2020-10 | ···················· | ·A·················· | ···················· | ···················· | ···················· | ···················· |
| AA | 1585 hCoV-19/USA/HI-H200496/2020\|EPI_ISL_753035\|2020-10 | ···················· | ·A·················· | ···················· | ···················· | ···················· | ···················· |
| AA | 1586 hCoV-19/USA/HI-H200497/2020\|EPI_ISL_753036\|2020-10 | ···················· | ·A·················· | ···················· | ···················· | ···················· | ···················· |
| AA | 1587 hCoV-19/USA/HI-H200498/2020\|EPI_ISL_753037\|2020-10 | ···················· | ·A·················· | ···················· | ···················· | ···················· | ···················· |
| AA | 1588 hCoV-19/USA/HI-H200499/2020\|EPI_ISL_753038\|2020-10 | ···················· | ·A·················· | ···················· | ···················· | ···················· | ···················· |
| AA | 1589 hCoV-19/USA/HI-H200500/2020\|EPI_ISL_753039\|2020-10 | ···················· | ···················· | ···················· | ···················· | ···················· | ···············A···· |
| AA | 1590 hCoV-19/USA/HI-H200501/2020\|EPI_ISL_753040\|2020-08 | ···················· | ·A·················· | ···················· | ···················· | ···················· | ···················· |
| AA | 1591 hCoV-19/USA/HI-H200502/2020\|EPI_ISL_753041\|2020-08 | ···················· | ·A·················· | ···················· | ···················· | ···················· | ···················· |
| AA | 1592 hCoV-19/USA/HI-H200503/2020\|EPI_ISL_753042\|2020-08 | ···················· | ·A·················· | ···················· | ···················· | ···················· | ···················· |
| AA | 1593 hCoV-19/USA/HI-H200504/2020\|EPI_ISL_753043\|2020-08 | ···················· | ·A·················· | ···················· | ···················· | ···················· | ···················· |
| AA | 1594 hCoV-19/USA/HI-H200505/2020\|EPI_ISL_753044\|2020-08 | ···················· | ·A·················· | ···················· | ···················· | ···················· | ···················· |
| AA | 1595 hCoV-19/USA/HI-H200507/2020\|EPI_ISL_753045\|2020-08 | ···················· | ·A·················· | ···················· | ···················· | ···················· | ···················· |
| AA | 1596 hCoV-19/USA/HI-H200508/2020\|EPI_ISL_753046\|2020-08 | ···················· | ·A·················· | ···················· | ···················· | ···················· | ···················· |
| AA | 1597 hCoV-19/USA/HI-H200509/2020\|EPI_ISL_753047\|2020-08 | ···················· | ·A·················· | ···················· | ···················· | ···················· | ···················· |
| AA | 1598 hCoV-19/USA/HI-H200510/2020\|EPI_ISL_753048\|2020-08 | ···················· | ·A·················· | ···················· | ···················· | ···················· | ···················· |
| AA | 1599 hCoV-19/USA/HI-H200513/2020\|EPI_ISL_753049\|2020-08 | ···················· | ·A·················· | ···················· | ···················· | ···················· | ···················· |
| AA | 1600 hCoV-19/USA/HI-H200514/2020\|EPI_ISL_753050\|2020-08 | ···················· | ·A·················· | ···················· | ···················· | ···················· | ···················· |
| AA | 1601 hCoV-19/USA/HI-H200516/2020\|EPI_ISL_753052\|2020-08 | ···················· | ·A·················· | ···················· | ···················· | ···················· | ···················· |
| AA | 1602 hCoV-19/USA/HI-H200519/2020\|EPI_ISL_753053\|2020-09 | ···················· | ·A·················· | ···················· | ···················· | ···················· | ···················· |

| | | | | | | | |
|---|---|---|---|---|---|---|---|
| AA | 1603 hCoV-19/USA/HI-H200520/2020\|EPI_ISL_753054\|2020-10 | ···················· | ···················· | ···················· | ···················· | ···················· | ···················· |
| AA | 1604 hCoV-19/USA/HI-H200521/2020\|EPI_ISL_753055\|2020-11 | ···················· | ·A·················· | ···················· | ···················· | ···················· | ···················· |
| AA | 1605 hCoV-19/USA/HI-H200522/2020\|EPI_ISL_753056\|2020-11 | ···················· | ·A·················· | ···················· | ···················· | ···················· | ···················· |
| AA | 1606 hCoV-19/USA/HI-H200523/2020\|EPI_ISL_753057\|2020-11 | ···················· | ·A·················· | ···················· | ···················· | ···················· | ···················· |
| AA | 1607 hCoV-19/USA/HI-H200525/2020\|EPI_ISL_753058\|2020-11 | ···················· | ·A·················· | ···················· | ···················· | ···················· | ···················· |
| AA | 1608 hCoV-19/USA/HI-H200526/2020\|EPI_ISL_753059\|2020-11 | ···················· | ·A·················· | ···················· | ···················· | ···················· | ···················· |
| AA | 1609 hCoV-19/England/204961745/2020\|EPI_ISL_754305\|2020 | ···················· | ···················· | ···················· | ···················· | ···················· | ···················· |
| AA | 1610 hCoV-19/England/204980829/2020\|EPI_ISL_754306\|2020 | ···················· | ···················· | ···················· | ···················· | ···················· | ···················· |
| AA | 1611 hCoV-19/England/204981688/2020\|EPI_ISL_754307\|2020 | ···················· | ···················· | ···················· | ···················· | ···················· | ···················· |
| AA | 1612 hCoV-19/England/204990734/2020\|EPI_ISL_754308\|2020 | ···················· | ···················· | ···················· | ···················· | ···················· | ···················· |
| AA | 1613 hCoV-19/England/205000435/2020\|EPI_ISL_754309\|2020 | ···················· | ···················· | ···················· | ···················· | ···················· | ···················· |
| AA | 1614 hCoV-19/England/205000486/2020\|EPI_ISL_754310\|2020 | ···················· | ···················· | ···················· | ···················· | ···················· | ···················· |
| AA | 1615 hCoV-19/England/205021213/2020\|EPI_ISL_754311\|2020 | ···················· | ···················· | ···················· | ···················· | ···················· | ···················· |
| AA | 1616 hCoV-19/England/205260444/2020\|EPI_ISL_754312\|2020 | ···················· | ···················· | ···················· | ···················· | ···················· | ···················· |
| AA | 1617 hCoV-19/England/205260445/2020\|EPI_ISL_754313\|2020 | ···················· | ···················· | ···················· | ···················· | ···················· | ···················· |
| AA | 1618 hCoV-19/England/205260446/2020\|EPI_ISL_754314\|2020 | ···················· | ···················· | ···················· | ···················· | ···················· | ···················· |
| AA | 1619 hCoV-19/England/205260447/2020\|EPI_ISL_754315\|2020 | ···················· | ···················· | ···················· | ···················· | ···················· | ···················· |
| AA | 1620 hCoV-19/England/205260458/2020\|EPI_ISL_754316\|2020 | ···················· | ···················· | ···················· | ···················· | ···················· | ···················· |
| AA | 1621 hCoV-19/England/205260460/2020\|EPI_ISL_754317\|2020 | ···················· | ···················· | ···················· | ···················· | ···················· | ···················· |
| AA | 1622 hCoV-19/England/205260462/2020\|EPI_ISL_754318\|2020 | ···················· | ···················· | ···················· | ···················· | ···················· | ···················· |
| AA | 1623 hCoV-19/England/205260463/2020\|EPI_ISL_754319\|2020 | ···················· | ···················· | ···················· | ···················· | ···················· | ···················· |
| AA | 1624 hCoV-19/England/205260465/2020\|EPI_ISL_754320\|2020 | ···················· | ···················· | ···················· | ···················· | ···················· | ···············A···· |
| AA | 1625 hCoV-19/England/205260466/2020\|EPI_ISL_754321\|2020 | ···················· | ···················· | ···················· | ···················· | ···················· | ···················· |
| AA | 1626 hCoV-19/England/205260469/2020\|EPI_ISL_754322\|2020 | ···················· | ···················· | ···················· | ···················· | ···················· | ···················· |
| AA | 1627 hCoV-19/England/205260472/2020\|EPI_ISL_754323\|2020 | ···················· | ···················· | ···················· | ···················· | ···················· | ···················· |
| AA | 1628 hCoV-19/England/205260473/2020\|EPI_ISL_754324\|2020 | ···················· | ···················· | ···················· | ···················· | ···················· | ···················· |
| AA | 1629 hCoV-19/England/205260476/2020\|EPI_ISL_754325\|2020 | ···················· | ···················· | ···················· | ···················· | ···················· | ···················· |
| AA | 1630 hCoV-19/England/205260860/2020\|EPI_ISL_754326\|2020 | ···················· | ···················· | ···················· | ···················· | ···················· | ···················· |
| AA | 1631 hCoV-19/England/205260861/2020\|EPI_ISL_754327\|2020 | ···················· | ···················· | ···················· | ···················· | ···················· | ···················· |
| AA | 1632 hCoV-19/England/205260862/2020\|EPI_ISL_754328\|2020 | ···················· | ···················· | ···················· | ···················· | ···················· | ···················· |
| AA | 1633 hCoV-19/England/205260872/2020\|EPI_ISL_754329\|2020 | ···················· | ···················· | ···················· | ···················· | ···················· | ···················· |
| AA | 1634 hCoV-19/England/205260873/2020\|EPI_ISL_754330\|2020 | ···················· | ·A·················· | ···················· | ···················· | ···················· | ··A················· |
| AA | 1635 hCoV-19/England/205260878/2020\|EPI_ISL_754331\|2020 | ···················· | ···················· | ···················· | ···················· | ···················· | ···················· |
| AA | 1636 hCoV-19/England/204881129/2020\|EPI_ISL_754332\|2020 | ···················· | ···················· | ···················· | ···················· | ···················· | ···················· |
| AA | 1637 hCoV-19/England/204881383/2020\|EPI_ISL_754333\|2020 | ···················· | ···················· | ···················· | ···················· | ···················· | ···················· |
| AA | 1638 hCoV-19/England/204881385/2020\|EPI_ISL_754334\|2020 | ···················· | ···················· | ···················· | ···················· | ···················· | ···················· |
| AA | 1639 hCoV-19/England/204890402/2020\|EPI_ISL_754335\|2020 | ···················· | ···················· | ···················· | ···················· | ···················· | ···················· |
| AA | 1640 hCoV-19/England/204890403/2020\|EPI_ISL_754336\|2020 | ···················· | ···················· | ···················· | ···················· | ···················· | ···················· |
| AA | 1641 hCoV-19/England/204890529/2020\|EPI_ISL_754337\|2020 | ···················· | ···················· | ···················· | ···················· | ···················· | ···················· |
| AA | 1642 hCoV-19/England/204890704/2020\|EPI_ISL_754338\|2020 | ···················· | ···················· | ···················· | ···················· | ···················· | ···················· |
| AA | 1643 hCoV-19/England/204900001/2020\|EPI_ISL_754339\|2020 | ···················· | ···················· | ···················· | ···················· | ···················· | ···················· |
| AA | 1644 hCoV-19/England/204900563/2020\|EPI_ISL_754340\|2020 | ···················· | ···················· | ···················· | ···················· | ···················· | ···················· |
| AA | 1645 hCoV-19/England/204920919/2020\|EPI_ISL_754341\|2020 | ···················· | ···················· | ···················· | ···················· | ···················· | ···················· |
| AA | 1646 hCoV-19/England/204941327/2020\|EPI_ISL_754342\|2020 | ···················· | ···················· | ···················· | ···················· | ···················· | ···················· |
| AA | 1647 hCoV-19/England/204941386/2020\|EPI_ISL_754343\|2020 | ···················· | ···················· | ···················· | ···················· | ···················· | ···················· |
| AA | 1648 hCoV-19/England/204941525/2020\|EPI_ISL_754344\|2020 | ···················· | ···················· | ···················· | ···················· | ···················· | ···················· |
| AA | 1649 hCoV-19/England/204941587/2020\|EPI_ISL_754345\|2020 | ···················· | ···················· | ···················· | ···················· | ···················· | ···················· |
| AA | 1650 hCoV-19/England/204960393/2020\|EPI_ISL_754346\|2020 | ···················· | ···················· | ···················· | ···················· | ···················· | ···················· |
| AA | 1651 hCoV-19/England/204960397/2020\|EPI_ISL_754347\|2020 | ···················· | ···················· | ···················· | ···················· | ···················· | ···················· |
| AA | 1652 hCoV-19/England/204961162/2020\|EPI_ISL_754348\|2020 | ···················· | ···················· | ···················· | ···················· | ···················· | ···················· |
| AA | 1653 hCoV-19/England/204980166/2020\|EPI_ISL_754349\|2020 | ···················· | ···················· | ···················· | ···················· | ···················· | ···················· |
| AA | 1654 hCoV-19/England/204980178/2020\|EPI_ISL_754350\|2020 | ···················· | ···················· | ···················· | ···················· | ···················· | ···················· |
| AA | 1655 hCoV-19/England/204980179/2020\|EPI_ISL_754351\|2020 | ···················· | ···················· | ···················· | ···················· | ···················· | ···················· |
| AA | 1656 hCoV-19/England/204980180/2020\|EPI_ISL_754352\|2020 | ···················· | ···················· | ···················· | ···················· | ···················· | ···················· |
| AA | 1657 hCoV-19/England/204980285/2020\|EPI_ISL_754353\|2020 | ···················· | ···················· | ···················· | ···················· | ···················· | ···················· |
| AA | 1658 hCoV-19/England/204980825/2020\|EPI_ISL_754354\|2020 | ···················· | ···················· | ···················· | ···················· | ···················· | ···················· |
| AA | 1659 hCoV-19/England/204981095/2020\|EPI_ISL_754355\|2020 | ···················· | ···················· | ···················· | ···················· | ···················· | ···················· |
| AA | 1660 hCoV-19/England/204990003/2020\|EPI_ISL_754356\|2020 | ···················· | ···················· | ···················· | ···················· | ···················· | ···················· |
| AA | 1661 hCoV-19/England/204991113/2020\|EPI_ISL_754357\|2020 | ···················· | ···················· | ···················· | ···················· | ···················· | ···················· |
| AA | 1662 hCoV-19/England/205000474/2020\|EPI_ISL_754358\|2020 | ···················· | ···················· | ···················· | ···················· | ···················· | ···················· |
| AA | 1663 hCoV-19/England/205000535/2020\|EPI_ISL_754359\|2020 | ···················· | ···················· | ···················· | ···················· | ···················· | ···················· |
| AA | 1664 hCoV-19/England/205000537/2020\|EPI_ISL_754360\|2020 | ···················· | ···················· | ···················· | ···················· | ···················· | ···················· |
| AA | 1665 hCoV-19/England/205021130/2020\|EPI_ISL_754361\|2020 | ···················· | ···················· | ···················· | ···················· | ···················· | ···················· |
| AA | 1666 hCoV-19/England/205260449/2020\|EPI_ISL_754362\|2020 | ···················· | ···················· | ···················· | ···················· | ···················· | ···················· |
| AA | 1667 hCoV-19/England/205260451/2020\|EPI_ISL_754363\|2020 | ···················· | ···················· | ···················· | ···················· | ···················· | ···················· |
| AA | 1668 hCoV-19/England/205260453/2020\|EPI_ISL_754364\|2020 | ···················· | ···················· | ···················· | ···················· | ···················· | ···················· |
| AA | 1669 hCoV-19/England/205260454/2020\|EPI_ISL_754365\|2020 | ···················· | ···················· | ···················· | ···················· | ···················· | ···················· |
| AA | 1670 hCoV-19/England/205260456/2020\|EPI_ISL_754366\|2020 | ···················· | ···················· | ···················· | ···················· | ···················· | ···················· |
| AA | 1671 hCoV-19/England/205260461/2020\|EPI_ISL_754367\|2020 | ···················· | ···················· | ···················· | ···················· | ···················· | ···················· |
| AA | 1672 hCoV-19/England/205260471/2020\|EPI_ISL_754368\|2020 | ···················· | ···················· | ···················· | ···················· | ···················· | ···················· |
| AA | 1673 hCoV-19/England/205260475/2020\|EPI_ISL_754369\|2020 | ···················· | ···················· | ···················· | ···················· | ···················· | ···················· |
| AA | 1674 hCoV-19/England/205260866/2020\|EPI_ISL_754370\|2020 | ···················· | ···················· | ···················· | ···················· | ···················· | ···················· |
| AA | 1675 hCoV-19/England/205260868/2020\|EPI_ISL_754371\|2020 | ···················· | ···················· | ···················· | ···················· | ···················· | ···················· |
| AA | 1676 hCoV-19/England/205260870/2020\|EPI_ISL_754372\|2020 | ···················· | ···················· | ···················· | ···················· | ···················· | ···················· |
| AA | 1677 hCoV-19/England/205260874/2020\|EPI_ISL_754373\|2020 | ···················· | ···················· | ···················· | ···················· | ···················· | ···················· |
| AA | 1678 hCoV-19/England/205260875/2020\|EPI_ISL_754374\|2020 | ···················· | ···················· | ···················· | ···················· | ···················· | ···················· |
| AA | 1679 hCoV-19/England/205260877/2020\|EPI_ISL_754375\|2020 | ···················· | ·A·················· | ···················· | ···················· | ···················· | ···················· |
| AA | 1680 hCoV-19/England/205261290/2020\|EPI_ISL_754376\|2020 | ···················· | ···················· | ···················· | ···················· | ···················· | ···················· |
| AA | 1681 hCoV-19/England/205261298/2020\|EPI_ISL_754377\|2020 | ···················· | ···················· | ···················· | ···················· | ···················· | ···················· |
| AA | 1682 hCoV-19/England/205260448/2020\|EPI_ISL_754378\|2020 | ···················· | ···················· | ···················· | ···················· | ···················· | ···················· |

| | | | | | | | | |
|---|---|---|---|---|---|---|---|---|
| AA | 1683 | hCoV-19/England/205260450/2020\|EPI_ISL_754379\|2020 | ···················· | ···················· | ···················· | ···················· | ···················· | ···················· |
| AA | 1684 | hCoV-19/England/205260452/2020\|EPI_ISL_754380\|2020 | ···················· | ···················· | ···················· | ···················· | ···················· | ···················· |
| AA | 1685 | hCoV-19/England/205260457/2020\|EPI_ISL_754381\|2020 | ···················· | ···················· | ···················· | ···················· | ···················· | ···················· |
| AA | 1686 | hCoV-19/England/205260459/2020\|EPI_ISL_754382\|2020 | ···················· | ···················· | ···················· | ···················· | ···················· | ···················· |
| AA | 1687 | hCoV-19/England/205260470/2020\|EPI_ISL_754383\|2020 | ···················· | ···················· | ···················· | ···················· | ···················· | ···················· |
| AA | 1688 | hCoV-19/England/205260474/2020\|EPI_ISL_754384\|2020 | ···················· | ···················· | ···················· | ···················· | ···················· | ···················· |
| AA | 1689 | hCoV-19/England/205260859/2020\|EPI_ISL_754385\|2020 | ···················· | ···················· | ···················· | ···················· | ···················· | ···················· |
| AA | 1690 | hCoV-19/England/205260864/2020\|EPI_ISL_754386\|2020 | ···················· | ···················· | ···················· | ···················· | ···················· | ···················· |
| AA | 1691 | hCoV-19/England/205260867/2020\|EPI_ISL_754387\|2020 | ···················· | ···················· | ···················· | ···················· | ···················· | ···················· |
| AA | 1692 | hCoV-19/USA/WY-WPHL-20156901/2020\|EPI_ISL_754388\|2 | ···················· | ···················· | ···················· | ···················· | ···················· | ···················· |
| AA | 1693 | hCoV-19/USA/WY-WPHL-20156911/2020\|EPI_ISL_754389\|2 | ···················· | ···················· | ···················· | ···················· | ···················· | ···················· |
| AA | 1694 | hCoV-19/New Zealand/20CV0717/2020\|EPI_ISL_755637\|2 | ···················· | ···················· | ···················· | ···················· | ···················· | ···················· |
| AA | 1695 | hCoV-19/Brazil/SP-874/2020\|EPI_ISL_755640\|2020-12- | ···················· | ···················· | ···················· | ···················· | ···················· | ···················· |
| AA | 1696 | hCoV-19/Brazil/SP-875/2020\|EPI_ISL_755641\|2020-12- | ···················· | ···················· | ···················· | ···················· | ···················· | ···················· |
| AA | 1697 | hCoV-19/Brazil/SP-876/2020\|EPI_ISL_755642\|2020-12- | ···················· | ···················· | ···················· | ···················· | ···················· | ···················· |
| AA | 1698 | hCoV-19/Brazil/SP-877/2020\|EPI_ISL_755643\|2020-12- | ···················· | ···················· | ···················· | ···················· | ···················· | ···················· |
| AA | 1699 | hCoV-19/Brazil/SP-878/2020\|EPI_ISL_755644\|2020-12- | ···················· | ····A··············· | ···················· | ···················· | ···················· | ···················· |
| AA | 1700 | hCoV-19/Brazil/SP-879/2020\|EPI_ISL_755645\|2020-12- | ···················· | ···················· | ···················· | ···················· | ···················· | ···················· |
| AA | 1701 | hCoV-19/Brazil/SP-880/2020\|EPI_ISL_755646\|2020-12- | ···················· | ···················· | ···················· | ···················· | ···················· | ···················· |
| AA | 1702 | hCoV-19/Brazil/SP-881/2020\|EPI_ISL_755647\|2020-12- | ···················· | ···················· | ···················· | ···················· | ···················· | ···················· |
| AA | 1703 | hCoV-19/Brazil/SP-882/2020\|EPI_ISL_755648\|2020-12- | ···················· | ···················· | ···················· | ···················· | ···················· | ···················· |
| AA | 1704 | hCoV-19/Brazil/SP-883/2020\|EPI_ISL_755649\|2020-12- | ···················· | ···················· | ···················· | ···················· | ···················· | ···················· |
| AA | 1705 | hCoV-19/Brazil/SP-884/2020\|EPI_ISL_755650\|2020-12- | ···················· | ···················· | ···················· | ···················· | ···················· | ···················· |
| AA | 1706 | hCoV-19/Brazil/SP-885/2020\|EPI_ISL_755651\|2020-12- | ···················· | ···················· | ···················· | ···················· | ···················· | ···················· |
| AA | 1707 | hCoV-19/Brazil/SP-886/2020\|EPI_ISL_755652\|2020-12- | ···················· | ···················· | ···················· | ···················· | ···················· | ···················· |
| AA | 1708 | hCoV-19/Brazil/SP-887/2020\|EPI_ISL_755653\|2020-12- | ···················· | ···················· | ···················· | ···················· | ···················· | ···················· |
| AA | 1709 | hCoV-19/Brazil/SP-888/2020\|EPI_ISL_755654\|2020-12- | ···················· | ···················· | ···················· | ···················· | ···················· | ···················· |
| AA | 1710 | hCoV-19/Brazil/SP-889/2020\|EPI_ISL_755655\|2020-12- | ···················· | ···················· | ···················· | ···················· | ···················· | ···················· |
| AA | 1711 | hCoV-19/Canada/ON-S1935/2020\|EPI_ISL_755656\|2020-1 | ···················· | ···················· | ···················· | ···················· | ···················· | ···················· |
| AA | 1712 | hCoV-19/Canada/ON-S1937/2020\|EPI_ISL_755658\|2020-1 | ···················· | ···················· | ···················· | ···················· | ···················· | ···················· |
| AA | 1713 | hCoV-19/Canada/ON-S1938/2020\|EPI_ISL_755659\|2020-1 | ···················· | ···················· | ···················· | ···················· | ···················· | ···················· |
| AA | 1714 | hCoV-19/Canada/ON-S1939/2020\|EPI_ISL_755660\|2020-1 | ···················· | ···················· | ···················· | ···················· | ···················· | ···················· |
| AA | 1715 | hCoV-19/Canada/ON-S1941/2020\|EPI_ISL_755661\|2020-1 | ···················· | ···················· | ···················· | ···················· | ···················· | ···················· |
| AA | 1716 | hCoV-19/Canada/ON-S1942/2020\|EPI_ISL_755662\|2020-1 | ···················· | ···················· | ···················· | ···················· | ···················· | ···················· |
| AA | 1717 | hCoV-19/Canada/ON-S1943/2020\|EPI_ISL_755663\|2020-1 | ···················· | ···················· | ···················· | ···················· | ···················· | ···················· |
| AA | 1718 | hCoV-19/Canada/ON-S1944/2020\|EPI_ISL_755664\|2020-1 | ···················· | ·A·················· | ···················· | ···················· | ···················· | ···A················ |
| AA | 1719 | hCoV-19/Canada/ON-S1945/2020\|EPI_ISL_755665\|2020-1 | ···················· | ···················· | ···················· | ···················· | ···················· | ···················· |
| AA | 1720 | hCoV-19/Canada/ON-S1946/2020\|EPI_ISL_755666\|2020-1 | ···················· | ·A·················· | ···················· | ···················· | ···················· | ···A················ |
| AA | 1721 | hCoV-19/Canada/ON-S1947/2020\|EPI_ISL_755667\|2020-1 | ···················· | ···················· | ···················· | ···················· | ···················· | ···················· |
| AA | 1722 | hCoV-19/Canada/ON-S1948/2020\|EPI_ISL_755668\|2020-1 | ···················· | ···················· | ···················· | ···················· | ···················· | ···················· |
| AA | 1723 | hCoV-19/Canada/ON-S1951/2020\|EPI_ISL_755671\|2020-1 | ···················· | ···················· | ···················· | ···················· | ···················· | ···················· |
| AA | 1724 | hCoV-19/Canada/ON-S1952/2020\|EPI_ISL_755672\|2020-1 | ···················· | ·A·················· | ···················· | ···················· | ···················· | ···A················ |
| AA | 1725 | hCoV-19/Canada/ON-S1954/2020\|EPI_ISL_755674\|2020-1 | ···················· | ·A·················· | ···················· | ···················· | ···················· | ···A················ |
| AA | 1726 | hCoV-19/Canada/ON-S1955/2020\|EPI_ISL_755675\|2020-1 | ···················· | ···················· | ···················· | ···················· | ···················· | ···················· |
| AA | 1727 | hCoV-19/Canada/ON-S1957/2020\|EPI_ISL_755676\|2020-1 | ···················· | ···················· | ···················· | ···················· | ···················· | ···················· |
| AA | 1728 | hCoV-19/Canada/ON-S1958/2020\|EPI_ISL_755677\|2020-1 | ···················· | ···················· | ···················· | ···················· | ···················· | ···················· |
| AA | 1729 | hCoV-19/Canada/ON-S1962/2020\|EPI_ISL_755678\|2020-1 | ···················· | ·A·················· | ···················· | ···················· | ···················· | ···A················ |
| AA | 1730 | hCoV-19/Canada/ON-S1963/2020\|EPI_ISL_755679\|2020-1 | ···················· | ·A·················· | ···················· | ···················· | ···················· | ···A················ |
| AA | 1731 | hCoV-19/Canada/ON-S1965/2020\|EPI_ISL_755681\|2020-1 | ···················· | ·A·················· | ···················· | ···················· | ···················· | ···A················ |
| AA | 1732 | hCoV-19/Canada/ON-S1966/2020\|EPI_ISL_755682\|2020-1 | ···················· | ·A·················· | ···················· | ···················· | ···················· | ···A················ |
| AA | 1733 | hCoV-19/Canada/ON-S1967/2020\|EPI_ISL_755683\|2020-1 | ···················· | ·A·················· | ···················· | ···················· | ···················· | ···A················ |
| AA | 1734 | hCoV-19/Canada/ON-S1968/2020\|EPI_ISL_755684\|2020-1 | ···················· | ·A·················· | ···················· | ···················· | ···················· | ···A················ |
| AA | 1735 | hCoV-19/Canada/ON-S1971/2020\|EPI_ISL_755685\|2020-1 | ···················· | ···················· | ···················· | ···················· | ···················· | ···················· |
| AA | 1736 | hCoV-19/Canada/ON-S1972/2020\|EPI_ISL_755686\|2020-1 | ···················· | ·A·················· | ···················· | ···················· | ···················· | ···A················ |
| AA | 1737 | hCoV-19/Canada/ON-S1973/2020\|EPI_ISL_755687\|2020-1 | ···················· | ·A·················· | ···················· | ···················· | ···················· | ···A················ |
| AA | 1738 | hCoV-19/Canada/ON-S1974/2020\|EPI_ISL_755688\|2020-1 | ···················· | ·A·················· | ···················· | ···················· | ···················· | ···A················ |
| AA | 1739 | hCoV-19/Canada/ON-S1975/2020\|EPI_ISL_755689\|2020-1 | ···················· | ·A·················· | ···················· | ···················· | ···················· | ···A················ |
| AA | 1740 | hCoV-19/Canada/ON-S1976/2020\|EPI_ISL_755690\|2020-1 | ···················· | ·A·················· | ···················· | ···················· | ···················· | ···A················ |
| AA | 1741 | hCoV-19/Canada/ON-S1977/2020\|EPI_ISL_755691\|2020-1 | ···················· | ·A·················· | ···················· | ···················· | ···················· | ···A················ |
| AA | 1742 | hCoV-19/Canada/ON-S1978/2020\|EPI_ISL_755692\|2020-1 | ···················· | ·A·················· | ···················· | ···················· | ···················· | ···A················ |
| AA | 1743 | hCoV-19/Canada/ON-S1979/2020\|EPI_ISL_755693\|2020-1 | ···················· | ···················· | ···················· | ···················· | ···················· | ···················· |
| AA | 1744 | hCoV-19/Canada/ON-S1981/2020\|EPI_ISL_755694\|2020-1 | ···················· | ···················· | ···················· | ···················· | ···················· | ···················· |
| AA | 1745 | hCoV-19/Canada/ON-S1982/2020\|EPI_ISL_755695\|2020-1 | ···················· | ·········A·········· | ···················· | ···················· | ···················· | ···················· |
| AA | 1746 | hCoV-19/Canada/ON-S1983/2020\|EPI_ISL_755696\|2020-1 | ···················· | ···················· | ···················· | ···················· | ···················· | ···················· |
| AA | 1747 | hCoV-19/Canada/ON-S1984/2020\|EPI_ISL_755697\|2020-1 | ···················· | ·A·················· | ···················· | ···················· | ···················· | ···A················ |
| AA | 1748 | hCoV-19/Canada/ON-S1985/2020\|EPI_ISL_755698\|2020-1 | ···················· | ·A·················· | ···················· | ···················· | ···················· | ···A················ |
| AA | 1749 | hCoV-19/Canada/ON-S1986/2020\|EPI_ISL_755699\|2020-1 | ···················· | ···················· | ···················· | ···················· | ···················· | ···················· |
| AA | 1750 | hCoV-19/England/MILK-C78298/2020\|EPI_ISL_756904\|20 | ···················· | ···················· | ···················· | ···················· | ···················· | ···················· |
| AA | 1751 | hCoV-19/England/MILK-C7BA60/2020\|EPI_ISL_756905\|20 | ···················· | ···················· | ···················· | ···················· | ···················· | ···················· |
| AA | 1752 | hCoV-19/England/QEUH-C7C258/2020\|EPI_ISL_756910\|20 | ···················· | ···················· | ···················· | ···················· | ···················· | ···················· |
| AA | 1753 | hCoV-19/England/MILK-C784B0/2020\|EPI_ISL_756915\|20 | ···················· | ·A·················· | ···················· | ···················· | ···················· | ···A················ |
| AA | 1754 | hCoV-19/England/QEUH-C7C23A/2020\|EPI_ISL_756971\|20 | ···················· | ···················· | ···················· | ···················· | ···················· | ···················· |
| AA | 1755 | hCoV-19/England/MILK-C789FD/2020\|EPI_ISL_756978\|20 | ···················· | ···················· | ···················· | ···················· | ···················· | ···················· |
| AA | 1756 | hCoV-19/England/MILK-C78483/2020\|EPI_ISL_756985\|20 | ···················· | ···················· | ···················· | ···················· | ···················· | ···················· |
| AA | 1757 | hCoV-19/England/MILK-C784FC/2020\|EPI_ISL_756988\|20 | ···················· | ···················· | ···················· | ···················· | ···················· | ···················· |
| AA | 1758 | hCoV-19/England/MILK-C7B541/2020\|EPI_ISL_756993\|20 | ···················· | ···················· | ···················· | ···················· | ···················· | ···················· |
| AA | 1759 | hCoV-19/England/MILK-C7B936/2020\|EPI_ISL_756994\|20 | ········T··········· | ·A·················· | ···················· | ···················· | ···················· | ···················· |
| AA | 1760 | hCoV-19/England/QEUH-C7C829/2020\|EPI_ISL_756998\|20 | ···················· | ···················· | ···················· | ···················· | ···················· | ···················· |
| AA | 1761 | hCoV-19/Denmark/DCGC-20462/2020\|EPI_ISL_759166\|202 | ···················· | ···················· | ···················· | ···················· | ···················· | ···················· |
| AA | 1762 | hCoV-19/Denmark/DCGC-20464/2020\|EPI_ISL_759167\|202 | ···················· | ···················· | ···················· | ···················· | ···················· | ···················· |

| | | | | | | | |
|---|---|---|---|---|---|---|---|
| AA | 1763 hCoV-19/Denmark/DCGC-20468/2020\|EPI_ISL_759170\|202 | .................... | .................... | .................... | .................... | .................... | .................... |
| AA | 1764 hCoV-19/Denmark/DCGC-20471/2020\|EPI_ISL_759171\|202 | .................... | .................... | .................... | .................... | .................... | .................... |
| AA | 1765 hCoV-19/Denmark/DCGC-20473/2020\|EPI_ISL_759172\|202 | .................... | .................... | .................... | .................... | .................... | .................... |
| AA | 1766 hCoV-19/Denmark/DCGC-20475/2020\|EPI_ISL_759173\|202 | .................... | .................... | .................... | .................... | .................... | .................... |
| AA | 1767 hCoV-19/Denmark/DCGC-20476/2020\|EPI_ISL_759174\|202 | .................... | .................... | .................... | .................... | .................... | .................... |
| AA | 1768 hCoV-19/Denmark/DCGC-20477/2020\|EPI_ISL_759175\|202 | .................... | .................... | .................... | .................... | .................... | .................... |
| AA | 1769 hCoV-19/Denmark/DCGC-20480/2020\|EPI_ISL_759176\|202 | .................... | .................... | .................... | .................... | .................... | .................... |
| AA | 1770 hCoV-19/Denmark/DCGC-20482/2020\|EPI_ISL_759177\|202 | .................... | .................... | .................... | .................... | .................... | .................... |
| AA | 1771 hCoV-19/Denmark/DCGC-20484/2020\|EPI_ISL_759178\|202 | .................... | .................... | .................... | .................... | .................... | .................... |
| AA | 1772 hCoV-19/Denmark/DCGC-20486/2020\|EPI_ISL_759179\|202 | .................... | .................... | .................... | .................... | .................... | .................... |
| AA | 1773 hCoV-19/Denmark/DCGC-20488/2020\|EPI_ISL_759180\|202 | .................... | .................... | .................... | .................... | .................... | .................... |
| AA | 1774 hCoV-19/Denmark/DCGC-20490/2020\|EPI_ISL_759181\|202 | .................... | .................... | .................... | .................... | .................... | .................... |
| AA | 1775 hCoV-19/Denmark/DCGC-20494/2020\|EPI_ISL_759182\|202 | .................... | .................... | .................... | .................... | .................... | .................... |
| AA | 1776 hCoV-19/Denmark/DCGC-20495/2020\|EPI_ISL_759183\|202 | .................... | .................... | .................... | .................... | .................... | .................... |
| AA | 1777 hCoV-19/Denmark/DCGC-20496/2020\|EPI_ISL_759184\|202 | .................... | .................... | .................... | .................... | .................... | .................... |
| AA | 1778 hCoV-19/Denmark/DCGC-20497/2020\|EPI_ISL_759185\|202 | .................... | .................... | .................... | .................... | .................... | .................... |
| AA | 1779 hCoV-19/Denmark/DCGC-20498/2020\|EPI_ISL_759186\|202 | .................... | .................... | .................... | .................... | .................... | .................... |
| AA | 1780 hCoV-19/Denmark/DCGC-20499/2020\|EPI_ISL_759187\|202 | .................... | .................... | .................... | .................... | .................... | .................... |
| AA | 1781 hCoV-19/Denmark/DCGC-20500/2020\|EPI_ISL_759188\|202 | .................... | .................... | .................... | .................... | .................... | .................... |
| AA | 1782 hCoV-19/Denmark/DCGC-20501/2020\|EPI_ISL_759189\|202 | .................... | .................... | .................... | .................... | .................... | .................... |
| AA | 1783 hCoV-19/Denmark/DCGC-20502/2020\|EPI_ISL_759190\|202 | .................... | .................... | .................... | .................... | .................... | .................... |
| AA | 1784 hCoV-19/Denmark/DCGC-20504/2020\|EPI_ISL_759191\|202 | .................... | .................... | .................... | .................... | .................... | .................... |
| AA | 1785 hCoV-19/Denmark/DCGC-20505/2020\|EPI_ISL_759192\|202 | .................... | .................... | .................... | .................... | .................... | .................... |
| AA | 1786 hCoV-19/Denmark/DCGC-20506/2020\|EPI_ISL_759193\|202 | .................... | .................... | .................... | .................... | .................... | .................... |
| AA | 1787 hCoV-19/Denmark/DCGC-20507/2020\|EPI_ISL_759194\|202 | .................... | .................... | .................... | .................... | .................... | .................... |
| AA | 1788 hCoV-19/Denmark/DCGC-20508/2020\|EPI_ISL_759195\|202 | .................... | .................... | .................... | .................... | .................... | .................... |
| AA | 1789 hCoV-19/Denmark/DCGC-20509/2020\|EPI_ISL_759196\|202 | .................... | .................... | .................... | .................... | .................... | .................... |
| AA | 1790 hCoV-19/Denmark/DCGC-20510/2020\|EPI_ISL_759197\|202 | .................... | .A.................. | .................... | .................... | .................... | .................... |
| AA | 1791 hCoV-19/Denmark/DCGC-20514/2020\|EPI_ISL_759198\|202 | .................... | .................... | .................... | .................... | .................... | .................... |
| AA | 1792 hCoV-19/Denmark/DCGC-20517/2020\|EPI_ISL_759199\|202 | .................... | .................... | .................... | .................... | .................... | .................... |
| AA | 1793 hCoV-19/USA/WA-UW-45062/2020\|EPI_ISL_747200\|2020-1 | .................... | .................... | .................... | .................... | .................... | .................... |
| AA | 1794 hCoV-19/USA/WA-UW-45072/2020\|EPI_ISL_747201\|2020-1 | .................... | .A.................. | .................... | .................... | .................... | .................... |
| AA | 1795 hCoV-19/USA/WA-UW-45075/2020\|EPI_ISL_747202\|2020-1 | .................... | .A.................. | .................... | .................... | .................... | .................... |
| AA | 1796 hCoV-19/USA/WA-UW-45076/2020\|EPI_ISL_747203\|2020-1 | .................... | .................... | .................... | .................... | .................... | .................... |
| AA | 1797 hCoV-19/USA/WA-UW-45077/2020\|EPI_ISL_747204\|2020-1 | .................... | .A.................. | .................... | .................... | .................... | ..A................. |
| AA | 1798 hCoV-19/USA/WA-UW-45078/2020\|EPI_ISL_747205\|2020-1 | .................... | .A.................. | .................... | .................... | .................... | .................... |
| AA | 1799 hCoV-19/USA/WA-UW-45088/2020\|EPI_ISL_747206\|2020-1 | .................... | .................... | .................... | .................... | .................... | .................... |
| AA | 1800 hCoV-19/USA/WA-UW-45089/2020\|EPI_ISL_747207\|2020-1 | .................... | .................... | .................... | .................... | .................... | .................... |
| AA | 1801 hCoV-19/USA/WA-UW-45090/2020\|EPI_ISL_747208\|2020-1 | .................... | .................... | .................... | .................... | .................... | .................... |
| AA | 1802 hCoV-19/USA/WA-UW-45091/2020\|EPI_ISL_747209\|2020-1 | .................... | .................... | .................... | .................... | .................... | .................... |
| AA | 1803 hCoV-19/USA/WA-UW-45094/2020\|EPI_ISL_747210\|2020-1 | .................... | .................... | .................... | .................... | .................... | .................... |
| AA | 1804 hCoV-19/USA/WA-UW-45095/2020\|EPI_ISL_747211\|2020-1 | .................... | .................... | .................... | .................... | .................... | .................... |
| AA | 1805 hCoV-19/USA/WA-UW-45096/2020\|EPI_ISL_747212\|2020-1 | .................... | .A.................. | .................... | .................... | .................... | .................... |
| AA | 1806 hCoV-19/USA/WA-UW-45097/2020\|EPI_ISL_747213\|2020-1 | .................... | .................... | .................... | .................... | .................... | .................... |
| AA | 1807 hCoV-19/USA/WA-UW-45099/2020\|EPI_ISL_747215\|2020-1 | .................... | .................... | .................... | .................... | .................... | .................... |
| AA | 1808 hCoV-19/USA/WA-UW-45104/2020\|EPI_ISL_747216\|2020-1 | .................... | .................... | .................... | .................... | .................... | .................... |
| AA | 1809 hCoV-19/USA/WA-UW-45106/2020\|EPI_ISL_747217\|2020-1 | .................... | .................... | .................... | .................... | .................... | .................... |
| AA | 1810 hCoV-19/USA/WA-UW-45107/2020\|EPI_ISL_747218\|2020-1 | .................... | .................... | .................... | .................... | .................... | .................... |
| AA | 1811 hCoV-19/USA/WA-UW-45110/2020\|EPI_ISL_747219\|2020-1 | .................... | .................... | .................... | .................... | .................... | .................... |
| AA | 1812 hCoV-19/Indonesia/JB-WJHL-ITB-R119840/2020\|EPI_ISL | .................... | .................... | .................... | .................... | .................... | .................... |
| AA | 1813 hCoV-19/Indonesia/JB-WJHL-ITB-R120975/2020\|EPI_ISL | .................... | .................... | .................... | .................... | .................... | .................... |
| AA | 1814 hCoV-19/South Korea/KDCA0004/2020\|EPI_ISL_747245\|2 | .................... | .................... | .................... | .................... | .................... | .................... |
| AA | 1815 hCoV-19/South Korea/KDCA0005/2020\|EPI_ISL_747246\|2 | .................... | .................... | .................... | .................... | .................... | .................... |
| AA | 1816 hCoV-19/South Korea/KDCA0006/2020\|EPI_ISL_747247\|2 | .................... | .................... | .................... | .................... | .................... | .................... |
| AA | 1817 hCoV-19/South Korea/KDCA0007/2020\|EPI_ISL_747248\|2 | .................... | .................... | .................... | .................... | .................... | .................... |
| AA | 1818 hCoV-19/South Korea/KDCA0008/2020\|EPI_ISL_747249\|2 | .................... | .................... | .................... | .................... | .................... | .................... |
| AA | 1819 hCoV-19/South Korea/KDCA0009/2020\|EPI_ISL_747250\|2 | .................... | .................... | .................... | .................... | .................... | .................... |
| AA | 1820 hCoV-19/South Korea/KDCA0010/2020\|EPI_ISL_747251\|2 | .................... | .................... | .................... | .................... | .................... | .................... |
| AA | 1821 hCoV-19/South Korea/KDCA0011/2020\|EPI_ISL_747252\|2 | .................... | .................... | .................... | .................... | .................... | .................... |
| AA | 1822 hCoV-19/South Korea/KDCA0012/2020\|EPI_ISL_747253\|2 | .................... | .................... | .................... | .................... | .................... | .................... |
| AA | 1823 hCoV-19/South Korea/KDCA0013/2020\|EPI_ISL_747254\|2 | .................... | .................... | .................... | .................... | .................... | .................... |
| AA | 1824 hCoV-19/South Korea/KDCA0015/2020\|EPI_ISL_747256\|2 | .................... | .................... | .................... | .................... | .................... | .................... |
| AA | 1825 hCoV-19/South Korea/KDCA0016/2020\|EPI_ISL_747257\|2 | .................... | .................... | .................... | .................... | .................... | .................... |
| AA | 1826 hCoV-19/South Korea/KDCA0017/2020\|EPI_ISL_747258\|2 | .................... | .................... | .................... | .................... | .................... | .................... |
| AA | 1827 hCoV-19/South Korea/KDCA0018/2020\|EPI_ISL_747259\|2 | .................... | .................... | .................... | .................... | .................... | .................... |
| AA | 1828 hCoV-19/Denmark/DCGC-18749/2020\|EPI_ISL_748532\|202 | .................... | .................... | .................... | .................... | .................... | .................... |
| AA | 1829 hCoV-19/Denmark/DCGC-18750/2020\|EPI_ISL_748533\|202 | .................... | .................... | .................... | .................... | .................... | .................... |
| AA | 1830 hCoV-19/Denmark/DCGC-18751/2020\|EPI_ISL_748534\|202 | .................... | .................... | .................... | .................... | .................... | .................... |
| AA | 1831 hCoV-19/Denmark/DCGC-18752/2020\|EPI_ISL_748535\|202 | .................... | .................... | .................... | .................... | .................... | .................... |
| AA | 1832 hCoV-19/Denmark/DCGC-18753/2020\|EPI_ISL_748536\|202 | .................... | .................... | .................... | .................... | .................... | .................... |
| AA | 1833 hCoV-19/Denmark/DCGC-18755/2020\|EPI_ISL_748537\|202 | .................... | .................... | .................... | .................... | .................... | .................... |
| AA | 1834 hCoV-19/Denmark/DCGC-18757/2020\|EPI_ISL_748538\|202 | .................... | .................... | .................... | .................... | .................... | .................... |
| AA | 1835 hCoV-19/Denmark/DCGC-18758/2020\|EPI_ISL_748539\|202 | .................... | .................... | .................... | .................... | .................... | .................... |
| AA | 1836 hCoV-19/Denmark/DCGC-18762/2020\|EPI_ISL_748540\|202 | .................... | .................... | .................... | .................... | .................... | .................... |
| AA | 1837 hCoV-19/Denmark/DCGC-18763/2020\|EPI_ISL_748541\|202 | .................... | .................... | .................... | .................... | .................... | .................... |
| AA | 1838 hCoV-19/Denmark/DCGC-18765/2020\|EPI_ISL_748542\|202 | .................... | .................... | .................... | .................... | .................... | .................... |
| AA | 1839 hCoV-19/Denmark/DCGC-18766/2020\|EPI_ISL_748543\|202 | .................... | .A.................. | .................... | .................... | .................... | ..A................. |
| AA | 1840 hCoV-19/Denmark/DCGC-18767/2020\|EPI_ISL_748544\|202 | .................... | .................... | .................... | .................... | .................... | .................... |
| AA | 1841 hCoV-19/Denmark/DCGC-18768/2020\|EPI_ISL_748545\|202 | .................... | .................... | .................... | .................... | .................... | .................... |
| AA | 1842 hCoV-19/Denmark/DCGC-18771/2020\|EPI_ISL_748546\|202 | .................... | .................... | .................... | .................... | .................... | .................... |

AA 1843 hCoV-19/Denmark/DCGC-18772/2020|EPI_ISL_748547|202 ···················· ···················· ···················· ···················· ···················· ····················
AA 1844 hCoV-19/Denmark/DCGC-18775/2020|EPI_ISL_748548|202 ···················· ···················· ···················· ···················· ···················· ····················
AA 1845 hCoV-19/Denmark/DCGC-18776/2020|EPI_ISL_748549|202 ···················· ···················· ···················· ···················· ···················· ····················
AA 1846 hCoV-19/Denmark/DCGC-18811/2020|EPI_ISL_748570|202 ···················· ···················· ···················· ···················· ···················· ····················
AA 1847 hCoV-19/Denmark/DCGC-18812/2020|EPI_ISL_748571|202 ···················· ···················· ···················· ···················· ···················· ····················
AA 1848 hCoV-19/Denmark/DCGC-18814/2020|EPI_ISL_748572|202 ···················· ···················· ···················· ···················· ···················· ····················
AA 1849 hCoV-19/Denmark/DCGC-18815/2020|EPI_ISL_748573|202 ···················· ···················· ···················· ···················· ···················· ····················
AA 1850 hCoV-19/Denmark/DCGC-18816/2020|EPI_ISL_748574|202 ···················· ···················· ···················· ···················· ···················· ····················
AA 1851 hCoV-19/Denmark/DCGC-18817/2020|EPI_ISL_748575|202 ···················· ···················· ···················· ···················· ···················· ····················
AA 1852 hCoV-19/Denmark/DCGC-18818/2020|EPI_ISL_748576|202 ···················· ···················· ···················· ···················· ···················· ····················
AA 1853 hCoV-19/Denmark/DCGC-18819/2020|EPI_ISL_748577|202 ···················· ···················· ···················· ···················· ···················· ····················
AA 1854 hCoV-19/Denmark/DCGC-18822/2020|EPI_ISL_748578|202 ···················· ···················· ···················· ···················· ···················· ····················
AA 1855 hCoV-19/Denmark/DCGC-18824/2020|EPI_ISL_748579|202 ···················· ···················· ···················· ···················· ···················· ····················
AA 1856 hCoV-19/Denmark/DCGC-18825/2020|EPI_ISL_748580|202 ···················· ···················· ···················· ···················· ···················· ····················
AA 1857 hCoV-19/Denmark/DCGC-18827/2020|EPI_ISL_748581|202 ···················· ···················· ···················· ···················· ···················· ····················
AA 1858 hCoV-19/Denmark/DCGC-18828/2020|EPI_ISL_748582|202 ···················· ···················· ···················· ···················· ···················· ····················
AA 1859 hCoV-19/Denmark/DCGC-18829/2020|EPI_ISL_748583|202 ···················· ···················· ···················· ···················· ···················· ····················
AA 1860 hCoV-19/Denmark/DCGC-18833/2020|EPI_ISL_748584|202 ···················· ···················· ···················· ···················· ···················· ····················
AA 1861 hCoV-19/Denmark/DCGC-18834/2020|EPI_ISL_748585|202 ···················· ···················· ···················· ···················· ···················· ····················
AA 1862 hCoV-19/Denmark/DCGC-18835/2020|EPI_ISL_748586|202 ···················· ···················· ···················· ···················· ···················· ····················
AA 1863 hCoV-19/Denmark/DCGC-18836/2020|EPI_ISL_748587|202 ···················· ···················· ···················· ···················· ···················· ····················
AA 1864 hCoV-19/Denmark/DCGC-18838/2020|EPI_ISL_748588|202 ···················· ···················· ···················· ···················· ···················· ····················
AA 1865 hCoV-19/Denmark/DCGC-18839/2020|EPI_ISL_748589|202 ···················· ···················· ···················· ···················· ···················· ····················
AA 1866 hCoV-19/Denmark/DCGC-18185/2020|EPI_ISL_749800|202 ···················· ···················· ···················· ···················· ···················· ····················
AA 1867 hCoV-19/Denmark/DCGC-18186/2020|EPI_ISL_749801|202 ···················· ···················· ···················· ···················· ···················· ····················
AA 1868 hCoV-19/Denmark/DCGC-18187/2020|EPI_ISL_749802|202 ···················· ···················· ···················· ···················· ···················· ····················
AA 1869 hCoV-19/Denmark/DCGC-18188/2020|EPI_ISL_749803|202 ···················· ···················· ···················· ···················· ···················· ····················
AA 1870 hCoV-19/Denmark/DCGC-18190/2020|EPI_ISL_749804|202 ···················· ···················· ···················· ···················· ···················· ····················
AA 1871 hCoV-19/Denmark/DCGC-18192/2020|EPI_ISL_749805|202 ···················· ···················· ···················· ···················· ···················· ····················
AA 1872 hCoV-19/Denmark/DCGC-18193/2020|EPI_ISL_749806|202 ···················· ···················· ···················· ···················· ···················· ····················
AA 1873 hCoV-19/Denmark/DCGC-18194/2020|EPI_ISL_749807|202 ···················· ···················· ···················· ···················· ···················· ····················
AA 1874 hCoV-19/Denmark/DCGC-18196/2020|EPI_ISL_749808|202 ···················· ···················· ···················· ···················· ···················· ····················
AA 1875 hCoV-19/Denmark/DCGC-18197/2020|EPI_ISL_749809|202 ···················· ···················· ···················· ···················· ···················· ····················
AA 1876 hCoV-19/Denmark/DCGC-18198/2020|EPI_ISL_749810|202 ···················· ···················· ···················· ···················· ···················· ····················
AA 1877 hCoV-19/Denmark/DCGC-18201/2020|EPI_ISL_749811|202 ···················· ···················· ···················· ···················· ···················· ····················
AA 1878 hCoV-19/Denmark/DCGC-18202/2020|EPI_ISL_749812|202 ···················· ···················· ···················· ···················· ···················· ····················
AA 1879 hCoV-19/Denmark/DCGC-18203/2020|EPI_ISL_749813|202 ···················· ···················· ···················· ···················· ···················· ····················
AA 1880 hCoV-19/Denmark/DCGC-18205/2020|EPI_ISL_749814|202 ···················· ···················· ···················· ···················· ···················· ····················
AA 1881 hCoV-19/Denmark/DCGC-18209/2020|EPI_ISL_749815|202 ···················· ···················· ···················· ···················· ···················· ····················
AA 1882 hCoV-19/Denmark/DCGC-18210/2020|EPI_ISL_749816|202 ···················· ·A·················· ···················· ···················· ···················· ····················
AA 1883 hCoV-19/Denmark/DCGC-18211/2020|EPI_ISL_749817|202 ···················· ···················· ···················· ···················· ···················· ····················
AA 1884 hCoV-19/Denmark/DCGC-18212/2020|EPI_ISL_749818|202 ···················· ···················· ···················· ···················· ···················· ····················
AA 1885 hCoV-19/Denmark/DCGC-18213/2020|EPI_ISL_749819|202 ···················· ···················· ···················· ···················· ···················· ····················
AA 1886 hCoV-19/Denmark/DCGC-18215/2020|EPI_ISL_749820|202 ···················· ···················· ···················· ···················· ···················· ····················
AA 1887 hCoV-19/Denmark/DCGC-18216/2020|EPI_ISL_749821|202 ···················· ···················· ···················· ···················· ···················· ····················
AA 1888 hCoV-19/Denmark/DCGC-18218/2020|EPI_ISL_749822|202 ···················· ···················· ···················· ···················· ···················· ····················
AA 1889 hCoV-19/Denmark/DCGC-18219/2020|EPI_ISL_749823|202 ···················· ···················· ···················· ···················· ···················· ····················
AA 1890 hCoV-19/Denmark/DCGC-18221/2020|EPI_ISL_749824|202 ···················· ···················· ···················· ···················· ···················· ····················
AA 1891 hCoV-19/Denmark/DCGC-18222/2020|EPI_ISL_749825|202 ···················· ···················· ···················· ···················· ···················· ····················
AA 1892 hCoV-19/Denmark/DCGC-18226/2020|EPI_ISL_749826|202 ···················· ···················· ···················· ···················· ···················· ····················
AA 1893 hCoV-19/Denmark/DCGC-18228/2020|EPI_ISL_749827|202 ···················· ···················· ···················· ···················· ···················· ····················
AA 1894 hCoV-19/Denmark/DCGC-18229/2020|EPI_ISL_749828|202 ···················· ···················· ···················· ···················· ···················· ····················
AA 1895 hCoV-19/Denmark/DCGC-18231/2020|EPI_ISL_749829|202 ···················· ···················· ···················· ···················· ···················· ····················
AA 1896 hCoV-19/Denmark/DCGC-18233/2020|EPI_ISL_749830|202 ···················· ···················· ···················· ···················· ···················· ····················
AA 1897 hCoV-19/Denmark/DCGC-18238/2020|EPI_ISL_749834|202 ···················· ···················· ···················· ···················· ···················· ····················
AA 1898 hCoV-19/Denmark/DCGC-18245/2020|EPI_ISL_749839|202 ···················· ···················· ···················· ···················· ······T············· ····················
AA 1899 hCoV-19/Denmark/DCGC-18246/2020|EPI_ISL_749840|202 ···················· ···················· ···················· ···················· ···················· ····················
AA 1900 hCoV-19/Denmark/DCGC-18247/2020|EPI_ISL_749841|202 ···················· ···················· ···················· ···················· ···················· ····················
AA 1901 hCoV-19/Denmark/DCGC-18248/2020|EPI_ISL_749842|202 ···················· ···················· ···················· ···················· ···················· ····················
AA 1902 hCoV-19/Denmark/DCGC-18254/2020|EPI_ISL_749843|202 ···················· ···················· ···················· ···················· ···················· ····················
AA 1903 hCoV-19/Denmark/DCGC-18255/2020|EPI_ISL_749844|202 ···················· ···················· ···················· ···················· ···················· ····················
AA 1904 hCoV-19/Denmark/DCGC-18257/2020|EPI_ISL_749845|202 ···················· ···················· ···················· ···················· ···················· ····················
AA 1905 hCoV-19/Denmark/DCGC-18258/2020|EPI_ISL_749846|202 ···················· ···················· ···················· ···················· ···················· ····················
AA 1906 hCoV-19/Denmark/DCGC-18259/2020|EPI_ISL_749847|202 ···················· ···················· ···················· ···················· ···················· ····················
AA 1907 hCoV-19/Denmark/DCGC-18262/2020|EPI_ISL_749848|202 ···················· ···················· ···················· ···················· ···················· ····················
AA 1908 hCoV-19/Denmark/DCGC-18263/2020|EPI_ISL_749849|202 ···················· ···················· ···················· ···················· ···················· ····················
AA 1909 hCoV-19/Denmark/DCGC-18265/2020|EPI_ISL_749850|202 ···················· ···················· ···················· ···················· ···················· ····················
AA 1910 hCoV-19/Denmark/DCGC-18266/2020|EPI_ISL_749851|202 ···················· ···················· ···················· ···················· ···················· ····················
AA 1911 hCoV-19/Denmark/DCGC-18267/2020|EPI_ISL_749852|202 ···················· ···················· ···················· ···················· ···················· ····················
AA 1912 hCoV-19/Denmark/DCGC-18269/2020|EPI_ISL_749853|202 ···················· ···················· ···················· ···················· ···················· ····················
AA 1913 hCoV-19/Denmark/DCGC-18271/2020|EPI_ISL_749854|202 ···················· ·A·················· ···················· ···················· ···················· ····················
AA 1914 hCoV-19/Denmark/DCGC-18272/2020|EPI_ISL_749855|202 ···················· ···················· ···················· ···················· ···················· ····················
AA 1915 hCoV-19/Denmark/DCGC-18274/2020|EPI_ISL_749856|202 ···················· ···················· ···················· ···················· ···················· ····················
AA 1916 hCoV-19/Denmark/DCGC-18277/2020|EPI_ISL_749857|202 ···················· ···················· ···················· ···················· ···················· ····················
AA 1917 hCoV-19/Denmark/DCGC-18278/2020|EPI_ISL_749858|202 ···················· ···················· ···················· ···················· ···················· ····················
AA 1918 hCoV-19/Denmark/DCGC-18280/2020|EPI_ISL_749859|202 ···················· ···················· ···················· ···················· ···················· ····················
AA 1919 hCoV-19/Denmark/DCGC-18281/2020|EPI_ISL_749860|202 ···················· ···················· ···················· ···················· ···················· ····················
AA 1920 hCoV-19/Denmark/DCGC-18282/2020|EPI_ISL_749861|202 ···················· ···················· ···················· ···················· ···················· ····················
AA 1921 hCoV-19/Denmark/DCGC-18283/2020|EPI_ISL_749862|202 ···················· ···················· ···················· ···················· ···················· ····················
AA 1922 hCoV-19/Denmark/DCGC-18284/2020|EPI_ISL_749863|202 ···················· ···················· ···················· ···················· ···················· ····················

| | | | | | | | | |
|---|---|---|---|---|---|---|---|---|
| AA | 1923 | hCoV-19/Denmark/DCGC-18285/2020\|EPI_ISL_749864\|202 | ···················· | ···················· | ····················· | ···················· | ···················· | ···················· |
| AA | 1924 | hCoV-19/Denmark/DCGC-18288/2020\|EPI_ISL_749865\|202 | ···················· | ···················· | ····················· | ···················· | ···················· | ···················· |
| AA | 1925 | hCoV-19/Denmark/DCGC-18289/2020\|EPI_ISL_749866\|202 | ···················· | ···················· | ····················· | ···················· | ···················· | ···················· |
| AA | 1926 | hCoV-19/Denmark/DCGC-18290/2020\|EPI_ISL_749867\|202 | ···················· | ···················· | ····················· | ···················· | ···················· | ···················· |
| AA | 1927 | hCoV-19/Denmark/DCGC-18293/2020\|EPI_ISL_749868\|202 | ···················· | ···················· | ····················· | ···················· | ···················· | ···················· |
| AA | 1928 | hCoV-19/Denmark/DCGC-18295/2020\|EPI_ISL_749869\|202 | ···················· | ···················· | ····················· | ···················· | ···················· | ···················· |
| AA | 1929 | hCoV-19/Denmark/DCGC-18297/2020\|EPI_ISL_749870\|202 | ···················· | ···················· | ····················· | ···················· | ···················· | ···················· |
| AA | 1930 | hCoV-19/Denmark/DCGC-18298/2020\|EPI_ISL_749871\|202 | ···················· | ···················· | ····················· | ···················· | ···················· | ···················· |
| AA | 1931 | hCoV-19/Denmark/DCGC-18299/2020\|EPI_ISL_749872\|202 | ···················· | ···················· | ····················· | ···················· | ···················· | ···················· |
| AA | 1932 | hCoV-19/Denmark/DCGC-18300/2020\|EPI_ISL_749873\|202 | ···················· | ···················· | ····················· | ···················· | ···················· | ···················· |
| AA | 1933 | hCoV-19/Denmark/DCGC-18301/2020\|EPI_ISL_749874\|202 | ···················· | ···················· | ····················· | ···················· | ···················· | ···················· |
| AA | 1934 | hCoV-19/Denmark/DCGC-18302/2020\|EPI_ISL_749875\|202 | ···················· | ···················· | ····················· | ···················· | ···················· | ···················· |
| AA | 1935 | hCoV-19/Denmark/DCGC-18303/2020\|EPI_ISL_749876\|202 | ···················· | ···················· | ····················· | ···················· | ···················· | ···················· |
| AA | 1936 | hCoV-19/Denmark/DCGC-18305/2020\|EPI_ISL_749877\|202 | ···················· | ···················· | ····················· | ···················· | ···················· | ···················· |
| AA | 1937 | hCoV-19/Denmark/DCGC-18306/2020\|EPI_ISL_749878\|202 | ···················· | ·A·················· | ····················· | ···················· | ···················· | ···················· |
| AA | 1938 | hCoV-19/Denmark/DCGC-18307/2020\|EPI_ISL_749879\|202 | ···················· | ···················· | ····················· | ···················· | ···················· | ···················· |
| AA | 1939 | hCoV-19/Denmark/DCGC-17170/2020\|EPI_ISL_750800\|202 | ···················· | ···················· | ····················· | ···················· | ···················· | ···················· |
| AA | 1940 | hCoV-19/Denmark/DCGC-17171/2020\|EPI_ISL_750801\|202 | ···················· | ···················· | ····················· | ···················· | ···················· | ···················· |
| AA | 1941 | hCoV-19/Denmark/DCGC-17174/2020\|EPI_ISL_750802\|202 | ···················· | ···················· | ····················· | ···················· | ···················· | ···················· |
| AA | 1942 | hCoV-19/Denmark/DCGC-17175/2020\|EPI_ISL_750803\|202 | ···················· | ···················· | ····················· | ···················· | ···················· | ···················· |
| AA | 1943 | hCoV-19/Denmark/DCGC-17177/2020\|EPI_ISL_750804\|202 | ···················· | ···················· | ····················· | ···················· | ···················· | ···················· |
| AA | 1944 | hCoV-19/Denmark/DCGC-17178/2020\|EPI_ISL_750805\|202 | ···················· | ···················· | ····················· | ···················· | ···················· | ···················· |
| AA | 1945 | hCoV-19/Denmark/DCGC-17179/2020\|EPI_ISL_750806\|202 | ···················· | ·A·················· | ····················· | ···················· | ···················· | ··A················· |
| AA | 1946 | hCoV-19/Denmark/DCGC-17180/2020\|EPI_ISL_750807\|202 | ···················· | ···················· | ····················· | ···················· | ···················· | ···················· |
| AA | 1947 | hCoV-19/Denmark/DCGC-17183/2020\|EPI_ISL_750808\|202 | ···················· | ···················· | ····················· | ···················· | ···················· | ···················· |
| AA | 1948 | hCoV-19/Denmark/DCGC-17184/2020\|EPI_ISL_750809\|202 | ···················· | ···················· | ····················· | ···················· | ···················· | ···················· |
| AA | 1949 | hCoV-19/Denmark/DCGC-17185/2020\|EPI_ISL_750810\|202 | ···················· | ···················· | ····················· | ···················· | ···················· | ···················· |
| AA | 1950 | hCoV-19/Denmark/DCGC-17187/2020\|EPI_ISL_750811\|202 | ···················· | ···················· | ····················· | ···················· | ···················· | ···················· |
| AA | 1951 | hCoV-19/Denmark/DCGC-17189/2020\|EPI_ISL_750812\|202 | ···················· | ·A·················· | ····················· | ···················· | ···················· | ···················· |
| AA | 1952 | hCoV-19/Denmark/DCGC-17190/2020\|EPI_ISL_750813\|202 | ···················· | ···················· | ····················· | ···················· | ···················· | ···················· |
| AA | 1953 | hCoV-19/Denmark/DCGC-17193/2020\|EPI_ISL_750814\|202 | ···················· | ···················· | ····················· | ···················· | ···················· | ···················· |
| AA | 1954 | hCoV-19/Denmark/DCGC-17194/2020\|EPI_ISL_750815\|202 | ···················· | ···················· | ····················· | ···················· | ···················· | ···················· |
| AA | 1955 | hCoV-19/Denmark/DCGC-17195/2020\|EPI_ISL_750816\|202 | ···················· | ···················· | ····················· | ···················· | ···················· | ···················· |
| AA | 1956 | hCoV-19/Denmark/DCGC-17196/2020\|EPI_ISL_750817\|202 | ···················· | ···················· | ····················· | ···················· | ···················· | ···················· |
| AA | 1957 | hCoV-19/Denmark/DCGC-17198/2020\|EPI_ISL_750818\|202 | ···················· | ···················· | ····················· | ···················· | ···················· | ···················· |
| AA | 1958 | hCoV-19/Denmark/DCGC-17200/2020\|EPI_ISL_750819\|202 | ···················· | ···················· | ····················· | ···················· | ···················· | ···················· |
| AA | 1959 | hCoV-19/Denmark/DCGC-17208/2020\|EPI_ISL_750826\|202 | ···················· | ···················· | ····················· | ···················· | ···················· | ···················· |
| AA | 1960 | hCoV-19/Denmark/DCGC-17209/2020\|EPI_ISL_750827\|202 | ···················· | ···················· | ····················· | ···················· | ···················· | ···················· |
| AA | 1961 | hCoV-19/Denmark/DCGC-17210/2020\|EPI_ISL_750828\|202 | ···················· | ···················· | ····················· | ···················· | ···················· | ···················· |
| AA | 1962 | hCoV-19/Denmark/DCGC-17211/2020\|EPI_ISL_750829\|202 | ···················· | ···················· | ····················· | ···················· | ···················· | ···················· |
| AA | 1963 | hCoV-19/Denmark/DCGC-17212/2020\|EPI_ISL_750830\|202 | ···················· | ···················· | ····················· | ···················· | ···················· | ···················· |
| AA | 1964 | hCoV-19/Denmark/DCGC-17213/2020\|EPI_ISL_750831\|202 | ···················· | ···················· | ····················· | ···················· | ···················· | ···················· |
| AA | 1965 | hCoV-19/Denmark/DCGC-17215/2020\|EPI_ISL_750832\|202 | ···················· | ···················· | ····················· | ···················· | ···················· | ···················· |
| AA | 1966 | hCoV-19/Denmark/DCGC-17216/2020\|EPI_ISL_750833\|202 | ···················· | ···················· | ····················· | ···················· | ···················· | ···················· |
| AA | 1967 | hCoV-19/Denmark/DCGC-17217/2020\|EPI_ISL_750834\|202 | ···················· | ···················· | ····················· | ···················· | ···················· | ···················· |
| AA | 1968 | hCoV-19/Denmark/DCGC-17219/2020\|EPI_ISL_750835\|202 | ···················· | ···················· | ····················· | ···················· | ···················· | ···················· |
| AA | 1969 | hCoV-19/Denmark/DCGC-17220/2020\|EPI_ISL_750836\|202 | ···················· | ···················· | ····················· | ···················· | ···················· | ···················· |
| AA | 1970 | hCoV-19/Denmark/DCGC-17223/2020\|EPI_ISL_750837\|202 | ···················· | ···················· | ····················· | ···················· | ···················· | ···················· |
| AA | 1971 | hCoV-19/Denmark/DCGC-17224/2020\|EPI_ISL_750838\|202 | ···················· | ···················· | ····················· | ···················· | ···················· | ···················· |
| AA | 1972 | hCoV-19/Denmark/DCGC-17225/2020\|EPI_ISL_750839\|202 | ···················· | ···················· | ····················· | ···················· | ···················· | ···················· |
| AA | 1973 | hCoV-19/Denmark/DCGC-17227/2020\|EPI_ISL_750840\|202 | ···················· | ···················· | ····················· | ···················· | ···················· | ···················· |
| AA | 1974 | hCoV-19/Denmark/DCGC-17228/2020\|EPI_ISL_750841\|202 | ···················· | ···················· | ····················· | ···················· | ···················· | ···················· |
| AA | 1975 | hCoV-19/Denmark/DCGC-17230/2020\|EPI_ISL_750842\|202 | ···················· | ···················· | ····················· | ···················· | ···················· | ···················· |
| AA | 1976 | hCoV-19/Denmark/DCGC-17232/2020\|EPI_ISL_750843\|202 | ···················· | ···················· | ····················· | ···················· | ···················· | ···················· |
| AA | 1977 | hCoV-19/Denmark/DCGC-17234/2020\|EPI_ISL_750844\|202 | ···················· | ···················· | ····················· | ···················· | ···················· | ···················· |
| AA | 1978 | hCoV-19/Denmark/DCGC-17236/2020\|EPI_ISL_750845\|202 | ···················· | ···················· | ····················· | ···················· | ···················· | ···················· |
| AA | 1979 | hCoV-19/Denmark/DCGC-17237/2020\|EPI_ISL_750846\|202 | ···················· | ···················· | ····················· | ···················· | ···················· | ···················· |
| AA | 1980 | hCoV-19/Denmark/DCGC-17238/2020\|EPI_ISL_750847\|202 | ···················· | ···················· | ····················· | ···················· | ···················· | ···················· |
| AA | 1981 | hCoV-19/Denmark/DCGC-17239/2020\|EPI_ISL_750848\|202 | ···················· | ···················· | ····················· | ···················· | ···················· | ···················· |
| AA | 1982 | hCoV-19/Denmark/DCGC-17241/2020\|EPI_ISL_750849\|202 | ···················· | ···················· | ····················· | ···················· | ···················· | ···················· |
| AA | 1983 | hCoV-19/Denmark/DCGC-17242/2020\|EPI_ISL_750850\|202 | ···················· | ···················· | ····················· | ···················· | ···················· | ···················· |
| AA | 1984 | hCoV-19/Denmark/DCGC-17244/2020\|EPI_ISL_750851\|202 | ···················· | ·A·················· | ····················· | ···················· | ···················· | ··A················· |
| AA | 1985 | hCoV-19/Denmark/DCGC-17245/2020\|EPI_ISL_750852\|202 | ···················· | ···················· | ····················· | ···················· | ···················· | ···················· |
| AA | 1986 | hCoV-19/Denmark/DCGC-17246/2020\|EPI_ISL_750853\|202 | ···················· | ···················· | ····················· | ···················· | ···················· | ···················· |
| AA | 1987 | hCoV-19/Denmark/DCGC-17248/2020\|EPI_ISL_750854\|202 | ···················· | ···················· | ····················· | ···················· | ···················· | ···················· |
| AA | 1988 | hCoV-19/Denmark/DCGC-17250/2020\|EPI_ISL_750855\|202 | ···················· | ·A·················· | ····················· | ···················· | ···················· | ··A················· |
| AA | 1989 | hCoV-19/Denmark/DCGC-17251/2020\|EPI_ISL_750856\|202 | ···················· | ···················· | ····················· | ···················· | ···················· | ···················· |
| AA | 1990 | hCoV-19/Denmark/DCGC-17252/2020\|EPI_ISL_750857\|202 | ···················· | ···················· | ····················· | ···················· | ···················· | ···················· |
| AA | 1991 | hCoV-19/Denmark/DCGC-17253/2020\|EPI_ISL_750858\|202 | ···················· | ···················· | ····················· | ···················· | ···················· | ···················· |
| AA | 1992 | hCoV-19/Denmark/DCGC-17254/2020\|EPI_ISL_750859\|202 | ···················· | ···················· | ····················· | ···················· | ···················· | ···················· |
| AA | 1993 | hCoV-19/Denmark/DCGC-17257/2020\|EPI_ISL_750860\|202 | ···················· | ···················· | ····················· | ···················· | ···················· | ···················· |
| AA | 1994 | hCoV-19/Denmark/DCGC-17259/2020\|EPI_ISL_750861\|202 | ···················· | ···················· | ····················· | ···················· | ···················· | ···················· |
| AA | 1995 | hCoV-19/USA/HI-H200527/2020\|EPI_ISL_753060\|2020-11 | ···················· | ·A·················· | ····················· | ···················· | ···················· | ···················· |
| AA | 1996 | hCoV-19/USA/HI-H200528/2020\|EPI_ISL_753061\|2020-11 | ···················· | ·A·················· | ····················· | ···················· | ···················· | ···················· |
| AA | 1997 | hCoV-19/USA/HI-H200529/2020\|EPI_ISL_753062\|2020-11 | ···················· | ·A·················· | ····················· | ···················· | ···················· | ···················· |
| AA | 1998 | hCoV-19/USA/HI-H200530/2020\|EPI_ISL_753063\|2020-10 | ···················· | ·A·················· | ····················· | ···················· | ···················· | ···················· |
| AA | 1999 | hCoV-19/USA/HI-H200532/2020\|EPI_ISL_753064\|2020-10 | ···················· | ·A·················· | ····················· | ···················· | ···················· | ···················· |
| AA | 2000 | hCoV-19/USA/HI-H200533/2020\|EPI_ISL_753065\|2020-10 | ···················· | ·A·················· | ····················· | ···················· | ···················· | ···················· |
| AA | 2001 | hCoV-19/USA/HI-H200534/2020\|EPI_ISL_753066\|2020-10 | ···················· | ·A·················· | ····················· | ···················· | ···················· | ···················· |
| AA | 2002 | hCoV-19/USA/HI-H200536/2020\|EPI_ISL_753067\|2020-10 | ···················· | ·A·················· | ····················· | ···················· | ···················· | ···················· |

AA 2003 hCoV-19/USA/HI-H200537/2020|EPI_ISL_753068|2020-10 ···················· ·A·················· ···················· ···················· ···················· ····················
AA 2004 hCoV-19/USA/HI-H200538/2020|EPI_ISL_753069|2020-10 ···················· ·A·················· ···················· ···················· ···················· ····················
AA 2005 hCoV-19/USA/HI-H200539/2020|EPI_ISL_753070|2020-11 ···················· ·A·················· ···················· ···················· ···················· ····················
AA 2006 hCoV-19/USA/HI-H200540/2020|EPI_ISL_753071|2020-11 ···················· ·A·················· ···················· ···················· ···················· ····················
AA 2007 hCoV-19/USA/HI-H200541/2020|EPI_ISL_753072|2020-11 ···················· ·A·················· ···················· ···················· ···················· ····················
AA 2008 hCoV-19/USA/HI-H200542/2020|EPI_ISL_753073|2020-10 ···················· ···················· ···················· ···················· ···················· ····················
AA 2009 hCoV-19/USA/HI-H200543/2020|EPI_ISL_753074|2020-11 ···················· ·A·················· ···················· ···················· ···················· ····················
AA 2010 hCoV-19/USA/HI-H200544/2020|EPI_ISL_753075|2020-11 ···················· ·A·················· ···················· ···················· ···················· ····················
AA 2011 hCoV-19/USA/HI-H200545/2020|EPI_ISL_753076|2020-11 ···················· ·A·················· ···················· ···················· ···················· ····················
AA 2012 hCoV-19/USA/HI-H200546/2020|EPI_ISL_753077|2020-11 ···················· ·A·················· ···················· ···················· ···················· ····················
AA 2013 hCoV-19/USA/HI-H200547/2020|EPI_ISL_753078|2020-11 ···················· ·A·················· ···················· ···················· ···················· ····················
AA 2014 hCoV-19/USA/HI-H200548/2020|EPI_ISL_753079|2020-11 ···················· ·A·················· ···················· ···················· ···················· ····················
AA 2015 hCoV-19/USA/HI-H200550/2020|EPI_ISL_753080|2020-10 ···················· ·A·················· ···················· ···················· ···················· ····················
AA 2016 hCoV-19/USA/HI-H200552/2020|EPI_ISL_753081|2020-10 ···················· ·A·················· ···················· ···················· ···················· ····················
AA 2017 hCoV-19/USA/HI-H200553/2020|EPI_ISL_753082|2020-10 ···················· ·A·················· ···················· ···················· ···················· ····················
AA 2018 hCoV-19/USA/HI-H200554/2020|EPI_ISL_753083|2020-10 ···················· ·A·················· ···················· ···················· ···················· ····················
AA 2019 hCoV-19/USA/HI-H200555/2020|EPI_ISL_753084|2020-10 ···················· ·A·················· ···················· ···················· ···················· ····················
AA 2020 hCoV-19/USA/HI-H200556/2020|EPI_ISL_753085|2020-10 ···················· ·A·················· ···················· ···················· ···················· ····················
AA 2021 hCoV-19/USA/HI-H200557/2020|EPI_ISL_753086|2020-10 ···················· ·A·················· ···················· ···················· ···················· ····················
AA 2022 hCoV-19/USA/HI-H200559/2020|EPI_ISL_753087|2020-10 ···················· ·A·················· ···················· ···················· ···················· ····················
AA 2023 hCoV-19/USA/HI-H200560/2020|EPI_ISL_753088|2020-10 ···················· ·A·················· ···················· ···················· ···················· ····················
AA 2024 hCoV-19/USA/HI-H200561/2020|EPI_ISL_753089|2020-10 ···················· ·A·················· ···················· ···················· ···················· ····················
AA 2025 hCoV-19/USA/HI-H200562/2020|EPI_ISL_753090|2020-10 ···················· ·A·················· ···················· ···················· ···················· ····················
AA 2026 hCoV-19/USA/HI-H200563/2020|EPI_ISL_753091|2020-10 ···················· ·A·················· ···················· ···················· ···················· ····················
AA 2027 hCoV-19/USA/HI-H200564/2020|EPI_ISL_753092|2020-10 ···················· ·A·················· ···················· ···················· ···················· ····················
AA 2028 hCoV-19/USA/HI-H200565/2020|EPI_ISL_753093|2020-10 ···················· ·A·················· ···················· ···················· ···················· ····················
AA 2029 hCoV-19/USA/HI-H200566/2020|EPI_ISL_753094|2020-10 ···················· ·A·················· ···················· ···················· ···················· ····················
AA 2030 hCoV-19/USA/HI-H200567/2020|EPI_ISL_753095|2020-10 ···················· ·A·················· ···················· ···················· ···················· ····················
AA 2031 hCoV-19/USA/HI-H200569/2020|EPI_ISL_753096|2020-10 ···················· ·A·················· ···················· ···················· ···················· ····················
AA 2032 hCoV-19/USA/HI-H200570/2020|EPI_ISL_753097|2020-10 ···················· ·A·················· ···················· ···················· ···················· ····················
AA 2033 hCoV-19/USA/HI-H200571/2020|EPI_ISL_753098|2020-10 ···················· ·A·················· ···················· ···················· ···················· ····················
AA 2034 hCoV-19/USA/HI-H200572/2020|EPI_ISL_753099|2020-10 ···················· ·A·················· ···················· ···················· ···················· ····················
AA 2035 hCoV-19/USA/CA-LACPHL-AF00048/2020|EPI_ISL_754390| ···················· ·A·················· ···················· ···················· ···················· ····················
AA 2036 hCoV-19/Chile/RM-126796/2020|EPI_ISL_754391|2020-0 ···················· ···················· ···················· ···················· ···················· ····················
AA 2037 hCoV-19/Chile/RM-170923/2020|EPI_ISL_754392|2020-0 ···················· ···················· ···················· ···················· ···················· ····················
AA 2038 hCoV-19/Chile/RM-191288/2020|EPI_ISL_754393|2020-0 ···················· ···················· ···················· ···················· ···················· ····················
AA 2039 hCoV-19/Denmark/DCGC-20590/2020|EPI_ISL_759240|202 ···················· ···················· ···················· ···················· ···················· ····················
AA 2040 hCoV-19/Denmark/DCGC-20592/2020|EPI_ISL_759241|202 ···················· ···················· ···················· ···················· ···················· ····················
AA 2041 hCoV-19/Denmark/DCGC-20593/2020|EPI_ISL_759242|202 ···················· ···················· ···················· ···················· ···················· ····················
AA 2042 hCoV-19/Denmark/DCGC-20594/2020|EPI_ISL_759243|202 ···················· ···················· ···················· ···················· ···················· ····················
AA 2043 hCoV-19/Denmark/DCGC-20595/2020|EPI_ISL_759244|202 ···················· ···················· ···················· ···················· ···················· ····················
AA 2044 hCoV-19/Denmark/DCGC-20596/2020|EPI_ISL_759245|202 ···················· ···················· ···················· ···················· ···················· ····················
AA 2045 hCoV-19/Denmark/DCGC-20597/2020|EPI_ISL_759246|202 ···················· ···················· ···················· ···················· ···················· ····················
AA 2046 hCoV-19/Denmark/DCGC-20598/2020|EPI_ISL_759247|202 ···················· ···················· ···················· ···················· ···················· ····················
AA 2047 hCoV-19/Denmark/DCGC-20601/2020|EPI_ISL_759248|202 ···················· ···················· ···················· ···················· ···················· ····················
AA 2048 hCoV-19/Denmark/DCGC-20603/2020|EPI_ISL_759249|202 ···················· ···················· ···················· ···················· ···················· ····················
AA 2049 hCoV-19/USA/WA-UW-45111/2020|EPI_ISL_747220|2020-1 ···················· ···················· ···················· ···················· ···················· ····················
AA 2050 hCoV-19/USA/WA-UW-45112/2020|EPI_ISL_747221|2020-1 ···················· ···················· ···················· ···················· ···················· ····················
AA 2051 hCoV-19/USA/WA-UW-45113/2020|EPI_ISL_747222|2020-1 ···················· ···················· ···················· ···················· ···················· ····················
AA 2052 hCoV-19/USA/WA-UW-45114/2020|EPI_ISL_747223|2020-1 ···················· ···················· ···················· ···················· ···················· ····················
AA 2053 hCoV-19/USA/WA-UW-45115/2020|EPI_ISL_747224|2020-1 ···················· ···················· ···················· ···················· ···················· ····················
AA 2054 hCoV-19/USA/WA-UW-45122/2020|EPI_ISL_747225|2020-1 ···················· ···················· ···················· ···················· ···················· ····················
AA 2055 hCoV-19/USA/WA-UW-45137/2020|EPI_ISL_747227|2020-1 ···················· ···················· ···················· ···················· ···················· ····················
AA 2056 hCoV-19/USA/WA-UW-45138/2020|EPI_ISL_747228|2020-1 ···················· ···················· ···················· ···················· ···················· ····················
AA 2057 hCoV-19/USA/WA-UW-45145/2020|EPI_ISL_747229|2020-1 ···················· ···················· ···················· ···················· ···················· ····················
AA 2058 hCoV-19/USA/WA-UW-45146/2020|EPI_ISL_747230|2020-1 ···················· ···················· ···················· ···················· ···················· ····················
AA 2059 hCoV-19/USA/WA-UW-45188/2020|EPI_ISL_747231|2020-1 ···················· ···················· ···················· ···················· ···················· ····················
AA 2060 hCoV-19/USA/WA-UW-45190/2020|EPI_ISL_747232|2020-1 ···················· ···················· ···················· ···················· ···················· ····················
AA 2061 hCoV-19/Indonesia/JK-NIHRD-IST002/2020|EPI_ISL_747 ···················· ···A················ ···················· ···················· ···················· ····················
AA 2062 hCoV-19/Indonesia/JK-NIHRD-PME1264/2020|EPI_ISL_74 ···················· ···················· ···················· ···················· ···················· ····················
AA 2063 hCoV-19/Indonesia/JB-WJHL-ITB-A02050/2020|EPI_ISL_ ···················· ···················· ···················· ···················· ···················· ···············A····
AA 2064 hCoV-19/Indonesia/JB-WJHL-ITB-A03267/2020|EPI_ISL_ ···················· ···················· ···················· ···················· ···················· ····················
AA 2065 hCoV-19/Indonesia/JB-WJHL-ITB-R104416/2020|EPI_ISL ···················· ···················· ···················· ···················· ···················· ····················
AA 2066 hCoV-19/Indonesia/JB-WJHL-ITB-R109969/2020|EPI_ISL ···················· ···················· ···················· ···················· ···················· ····················
AA 2067 hCoV-19/South Korea/KDCA0019/2020|EPI_ISL_747260|2 ···················· ···················· ···················· ···················· ···················· ····················
AA 2068 hCoV-19/South Korea/KDCA0020/2020|EPI_ISL_747261|2 ···················· ···················· ···················· ···················· ···················· ····················
AA 2069 hCoV-19/South Korea/KDCA0021/2020|EPI_ISL_747262|2 ···················· ···················· ···················· ···················· ···················· ····················
AA 2070 hCoV-19/South Korea/KDCA0024/2020|EPI_ISL_747263|2 ···················· ···················· ···················· ···················· ···················· ····················
AA 2071 hCoV-19/South Korea/KDCA0025/2020|EPI_ISL_747264|2 ···················· ···················· ···················· ···················· ···················· ····················
AA 2072 hCoV-19/South Korea/KDCA0026/2020|EPI_ISL_747265|2 ···················· ···················· ···················· ···················· ···················· ····················
AA 2073 hCoV-19/South Korea/KDCA0027/2020|EPI_ISL_747266|2 ···················· ···················· ···················· ···················· ···················· ····················
AA 2074 hCoV-19/South Korea/KDCA0028/2020|EPI_ISL_747267|2 ···················· ···················· ···················· ···················· ···················· ····················
AA 2075 hCoV-19/South Korea/KDCA0029/2020|EPI_ISL_747268|2 ···················· ···················· ···················· ···················· ···················· ····················
AA 2076 hCoV-19/South Korea/KDCA0030/2020|EPI_ISL_747269|2 ···················· ···················· ···················· ···················· ···················· ····················
AA 2077 hCoV-19/South Korea/KDCA0031/2020|EPI_ISL_747270|2 ···················· ···················· ···················· ···················· ···················· ····················
AA 2078 hCoV-19/South Korea/KDCA0032/2020|EPI_ISL_747271|2 ···················· ···················· ···················· ···················· ···················· ····················
AA 2079 hCoV-19/South Korea/KDCA0033/2020|EPI_ISL_747272|2 ···················· ···················· ···················· ···················· ···················· ····················
AA 2080 hCoV-19/South Korea/KDCA0034/2020|EPI_ISL_747273|2 ···················· ···················· ···················· ···················· ···················· ····················
AA 2081 hCoV-19/South Korea/KDCA0035/2020|EPI_ISL_747274|2 ···················· ···················· ···················· ···················· ···················· ····················
AA 2082 hCoV-19/South Korea/KDCA0036/2020|EPI_ISL_747275|2 ···················· ···················· ···················· ···················· ···················· ····················

| | | | | | | | | |
|---|---|---|---|---|---|---|---|---|
| AA | 2083 | hCoV-19/South Korea/KDCA0037/2020\|EPI_ISL_747276\|2 | ···················· | ···················· | ····················· | ···················· | ···················· | ···················· |
| AA | 2084 | hCoV-19/South Korea/KDCA0038/2020\|EPI_ISL_747277\|2 | ···················· | ···················· | ····················· | ···················· | ···················· | ···················· |
| AA | 2085 | hCoV-19/South Korea/KDCA0039/2020\|EPI_ISL_747278\|2 | ···················· | ···················· | ····················· | ···················· | ···················· | ···················· |
| AA | 2086 | hCoV-19/South Korea/KDCA0040/2020\|EPI_ISL_747279\|2 | ···················· | ···················· | ····················· | ···················· | ···················· | ···················· |
| AA | 2087 | hCoV-19/South Korea/KDCA0041/2020\|EPI_ISL_747280\|2 | ···················· | ···················· | ····················· | ···················· | ···················· | ···················· |
| AA | 2088 | hCoV-19/South Korea/KDCA0042/2020\|EPI_ISL_747281\|2 | ···················· | ···················· | ···········T········· | ···················· | ···················· | ···················· |
| AA | 2089 | hCoV-19/Bangladesh/DNAS-Jakir/2020\|EPI_ISL_747283\| | ···················· | ···················· | ····················· | ···················· | ···················· | ···················· |
| AA | 2090 | hCoV-19/South Korea/KDCA0043/2020\|EPI_ISL_747284\|2 | ···················· | ···················· | ····················· | ···················· | ···················· | ···················· |
| AA | 2091 | hCoV-19/South Korea/KDCA0044/2020\|EPI_ISL_747285\|2 | ···················· | ···················· | ····················· | ···················· | ···················· | ···················· |
| AA | 2092 | hCoV-19/South Korea/KDCA0045/2020\|EPI_ISL_747286\|2 | ···················· | ···················· | ····················· | ···················· | ···················· | ···················· |
| AA | 2093 | hCoV-19/South Korea/KDCA0046/2020\|EPI_ISL_747287\|2 | ···················· | ···················· | ····················· | ···················· | ···················· | ···················· |
| AA | 2094 | hCoV-19/South Korea/KDCA0048/2020\|EPI_ISL_747289\|2 | ···················· | ···················· | ····················· | ···················· | ···················· | ···················· |
| AA | 2095 | hCoV-19/South Korea/KDCA0049/2020\|EPI_ISL_747290\|2 | ···················· | ···················· | ····················· | ···················· | ···················· | ···················· |
| AA | 2096 | hCoV-19/South Korea/KDCA0050/2020\|EPI_ISL_747291\|2 | ···················· | ···················· | ····················· | ···················· | ···················· | ···················· |
| AA | 2097 | hCoV-19/South Korea/KDCA0051/2020\|EPI_ISL_747292\|2 | ···················· | ···················· | ····················· | ···················· | ···················· | ···················· |
| AA | 2098 | hCoV-19/South Korea/KDCA0052/2020\|EPI_ISL_747293\|2 | ···················· | ···················· | ····················· | ···················· | ···················· | ···················· |
| AA | 2099 | hCoV-19/South Korea/KDCA0053/2020\|EPI_ISL_747294\|2 | ···················· | ···················· | ····················· | ···················· | ···················· | ···················· |
| AA | 2100 | hCoV-19/South Korea/KDCA0054/2020\|EPI_ISL_747295\|2 | ···················· | ···················· | ····················· | ···················· | ···················· | ···················· |
| AA | 2101 | hCoV-19/South Korea/KDCA0055/2020\|EPI_ISL_747296\|2 | ···················· | ···················· | ····················· | ···················· | ···················· | ···················· |
| AA | 2102 | hCoV-19/South Korea/KDCA0056/2020\|EPI_ISL_747297\|2 | ···················· | ···················· | ····················· | ···················· | ···················· | ···················· |
| AA | 2103 | hCoV-19/South Korea/KDCA0057/2020\|EPI_ISL_747298\|2 | ···················· | ···················· | ····················· | ···················· | ···················· | ···················· |
| AA | 2104 | hCoV-19/South Korea/KDCA0058/2020\|EPI_ISL_747299\|2 | ···················· | ···················· | ····················· | ···················· | ···················· | ···················· |
| AA | 2105 | hCoV-19/Denmark/DCGC-18777/2020\|EPI_ISL_748550\|202 | ···················· | ···················· | ····················· | ···················· | ···················· | ···················· |
| AA | 2106 | hCoV-19/Denmark/DCGC-18779/2020\|EPI_ISL_748551\|202 | ···················· | ···················· | ····················· | ···················· | ···················· | ···················· |
| AA | 2107 | hCoV-19/Denmark/DCGC-18781/2020\|EPI_ISL_748552\|202 | ···················· | ···················· | ····················· | ···················· | ···················· | ···················· |
| AA | 2108 | hCoV-19/Denmark/DCGC-18784/2020\|EPI_ISL_748553\|202 | ···················· | ···················· | ····················· | ···················· | ···················· | ···················· |
| AA | 2109 | hCoV-19/Denmark/DCGC-18785/2020\|EPI_ISL_748554\|202 | ···················· | ···················· | ····················· | ···················· | ···················· | ···················· |
| AA | 2110 | hCoV-19/Denmark/DCGC-18786/2020\|EPI_ISL_748555\|202 | ···················· | ···················· | ····················· | ···················· | ···················· | ···················· |
| AA | 2111 | hCoV-19/Denmark/DCGC-18788/2020\|EPI_ISL_748556\|202 | ···················· | ···················· | ····················· | ···················· | ···················· | ···················· |
| AA | 2112 | hCoV-19/Denmark/DCGC-18789/2020\|EPI_ISL_748557\|202 | ···················· | ···················· | ····················· | ···················· | ···················· | ···················· |
| AA | 2113 | hCoV-19/Denmark/DCGC-18790/2020\|EPI_ISL_748558\|202 | ···················· | ···················· | ····················· | ···················· | ···················· | ···················· |
| AA | 2114 | hCoV-19/Denmark/DCGC-18793/2020\|EPI_ISL_748559\|202 | ···················· | ···················· | ····················· | ···················· | ···················· | ···················· |
| AA | 2115 | hCoV-19/Denmark/DCGC-18795/2020\|EPI_ISL_748560\|202 | ···················· | ···················· | ····················· | ···················· | ···················· | ···················· |
| AA | 2116 | hCoV-19/Denmark/DCGC-18798/2020\|EPI_ISL_748561\|202 | ···················· | ···················· | ····················· | ···················· | ···················· | ···················· |
| AA | 2117 | hCoV-19/Denmark/DCGC-18799/2020\|EPI_ISL_748562\|202 | ···················· | ···················· | ····················· | ···················· | ···················· | ···················· |
| AA | 2118 | hCoV-19/Denmark/DCGC-18801/2020\|EPI_ISL_748563\|202 | ···················· | ···················· | ····················· | ···················· | ···················· | ···················· |
| AA | 2119 | hCoV-19/Denmark/DCGC-18802/2020\|EPI_ISL_748564\|202 | ···················· | ···················· | ····················· | ···················· | ···················· | ···················· |
| AA | 2120 | hCoV-19/Denmark/DCGC-18803/2020\|EPI_ISL_748565\|202 | ···················· | ···················· | ····················· | ···················· | ···················· | ···················· |
| AA | 2121 | hCoV-19/Denmark/DCGC-18804/2020\|EPI_ISL_748566\|202 | ···················· | ···················· | ····················· | ···················· | ···················· | ···················· |
| AA | 2122 | hCoV-19/Denmark/DCGC-18805/2020\|EPI_ISL_748567\|202 | ···················· | ···················· | ····················· | ···················· | ···················· | ···················· |
| AA | 2123 | hCoV-19/Denmark/DCGC-18807/2020\|EPI_ISL_748568\|202 | ···················· | ···················· | ····················· | ···················· | ···················· | ···················· |
| AA | 2124 | hCoV-19/Denmark/DCGC-18809/2020\|EPI_ISL_748569\|202 | ···················· | ·A·················· | ····················· | ···················· | ···················· | ···A················ |
| AA | 2125 | hCoV-19/Denmark/DCGC-18841/2020\|EPI_ISL_748590\|202 | ···················· | ···················· | ····················· | ···················· | ···················· | ···················· |
| AA | 2126 | hCoV-19/Denmark/DCGC-18842/2020\|EPI_ISL_748591\|202 | ···················· | ···················· | ····················· | ···················· | ···················· | ···················· |
| AA | 2127 | hCoV-19/Denmark/DCGC-18843/2020\|EPI_ISL_748592\|202 | ···················· | ·A·················· | ····················· | ···················· | ···················· | ···A················ |
| AA | 2128 | hCoV-19/Denmark/DCGC-18845/2020\|EPI_ISL_748593\|202 | ···················· | ···················· | ····················· | ···················· | ···················· | ···················· |
| AA | 2129 | hCoV-19/Denmark/DCGC-18847/2020\|EPI_ISL_748594\|202 | ···················· | ···················· | ····················· | ···················· | ···················· | ···················· |
| AA | 2130 | hCoV-19/Denmark/DCGC-18848/2020\|EPI_ISL_748595\|202 | ···················· | ·A·················· | ····················· | ···················· | ···················· | ···A················ |
| AA | 2131 | hCoV-19/Denmark/DCGC-18849/2020\|EPI_ISL_748596\|202 | ···················· | ···················· | ····················· | ···················· | ···················· | ···················· |
| AA | 2132 | hCoV-19/Denmark/DCGC-18850/2020\|EPI_ISL_748597\|202 | ···················· | ···················· | ····················· | ···················· | ···················· | ···················· |
| AA | 2133 | hCoV-19/Denmark/DCGC-18853/2020\|EPI_ISL_748598\|202 | ···················· | ···················· | ····················· | ···················· | ···················· | ···················· |
| AA | 2134 | hCoV-19/Denmark/DCGC-18855/2020\|EPI_ISL_748599\|202 | ···················· | ···················· | ····················· | ···················· | ···················· | ···················· |
| AA | 2135 | hCoV-19/Denmark/DCGC-19565/2020\|EPI_ISL_749030\|202 | ···················· | ···················· | ····················· | ···················· | ···················· | ···················· |
| AA | 2136 | hCoV-19/Denmark/DCGC-19567/2020\|EPI_ISL_749031\|202 | ···················· | ···················· | ····················· | ···················· | ···················· | ···················· |
| AA | 2137 | hCoV-19/Denmark/DCGC-19568/2020\|EPI_ISL_749032\|202 | ···················· | ···················· | ····················· | ···················· | ···················· | ···················· |
| AA | 2138 | hCoV-19/Denmark/DCGC-19569/2020\|EPI_ISL_749033\|202 | ···················· | ···················· | ····················· | ···················· | ···················· | ···················· |
| AA | 2139 | hCoV-19/Denmark/DCGC-19570/2020\|EPI_ISL_749034\|202 | ···················· | ·A·················· | ····················· | ···················· | ···················· | ···················· |
| AA | 2140 | hCoV-19/Denmark/DCGC-19571/2020\|EPI_ISL_749035\|202 | ···················· | ···················· | ····················· | ···················· | ···················· | ···················· |
| AA | 2141 | hCoV-19/Denmark/DCGC-19572/2020\|EPI_ISL_749037\|202 | ···················· | ···················· | ····················· | ···················· | ···················· | ···················· |
| AA | 2142 | hCoV-19/Denmark/DCGC-19574/2020\|EPI_ISL_749038\|202 | ···················· | ···················· | ····················· | ···················· | ···················· | ···················· |
| AA | 2143 | hCoV-19/Denmark/DCGC-19578/2020\|EPI_ISL_749039\|202 | ···················· | ···················· | ····················· | ···················· | ···················· | ···················· |
| AA | 2144 | hCoV-19/Denmark/DCGC-18308/2020\|EPI_ISL_749880\|202 | ···················· | ···················· | ····················· | ···················· | ···················· | ···················· |
| AA | 2145 | hCoV-19/Denmark/DCGC-18309/2020\|EPI_ISL_749881\|202 | ···················· | ···················· | ····················· | ···················· | ···················· | ···················· |
| AA | 2146 | hCoV-19/Denmark/DCGC-18310/2020\|EPI_ISL_749882\|202 | ···················· | ···················· | ····················· | ···················· | ···················· | ···················· |
| AA | 2147 | hCoV-19/Denmark/DCGC-18311/2020\|EPI_ISL_749883\|202 | ···················· | ·A·················· | ····················· | ···················· | ···················· | ···A················ |
| AA | 2148 | hCoV-19/Denmark/DCGC-18315/2020\|EPI_ISL_749884\|202 | ···················· | ···················· | ····················· | ···················· | ···················· | ···················· |
| AA | 2149 | hCoV-19/Denmark/DCGC-18317/2020\|EPI_ISL_749885\|202 | ···················· | ···················· | ····················· | ···················· | ···················· | ···················· |
| AA | 2150 | hCoV-19/Denmark/DCGC-18318/2020\|EPI_ISL_749886\|202 | ···················· | ···················· | ····················· | ···················· | ···················· | ···················· |
| AA | 2151 | hCoV-19/Denmark/DCGC-18321/2020\|EPI_ISL_749887\|202 | ···················· | ···················· | ····················· | ···················· | ···················· | ···················· |
| AA | 2152 | hCoV-19/Denmark/DCGC-18322/2020\|EPI_ISL_749888\|202 | ···················· | ···················· | ····················· | ···················· | ···················· | ···················· |
| AA | 2153 | hCoV-19/Denmark/DCGC-18325/2020\|EPI_ISL_749889\|202 | ···················· | ···················· | ····················· | ···················· | ···················· | ···················· |
| AA | 2154 | hCoV-19/Denmark/DCGC-18327/2020\|EPI_ISL_749890\|202 | ···················· | ···················· | ····················· | ···················· | ···················· | ···················· |
| AA | 2155 | hCoV-19/Denmark/DCGC-18329/2020\|EPI_ISL_749891\|202 | ···················· | ···················· | ····················· | ···················· | ···················· | ···················· |
| AA | 2156 | hCoV-19/Denmark/DCGC-18330/2020\|EPI_ISL_749892\|202 | ···················· | ···················· | ····················· | ···················· | ···················· | ···················· |
| AA | 2157 | hCoV-19/Denmark/DCGC-18331/2020\|EPI_ISL_749893\|202 | ···················· | ···················· | ····················· | ···················· | ···················· | ···················· |
| AA | 2158 | hCoV-19/Denmark/DCGC-18332/2020\|EPI_ISL_749894\|202 | ···················· | ···················· | ····················· | ···················· | ···················· | ···················· |
| AA | 2159 | hCoV-19/Denmark/DCGC-18334/2020\|EPI_ISL_749895\|202 | ···················· | ···················· | ····················· | ···················· | ···················· | ···················· |
| AA | 2160 | hCoV-19/Denmark/DCGC-18335/2020\|EPI_ISL_749896\|202 | ···················· | ···················· | ····················· | ···················· | ···················· | ···················· |
| AA | 2161 | hCoV-19/Denmark/DCGC-18336/2020\|EPI_ISL_749897\|202 | ···················· | ···················· | ····················· | ···················· | ···················· | ···················· |
| AA | 2162 | hCoV-19/Denmark/DCGC-18338/2020\|EPI_ISL_749898\|202 | ···················· | ···················· | ····················· | ···················· | ···················· | ···················· |

| | | | | | | | | |
|---|---|---|---|---|---|---|---|---|
| AA | 2163 | hCoV-19/Denmark/DCGC-18340/2020\|EPI_ISL_749899\|202 | ···················· | ···················· | ···················· | ···················· | ···················· | ···················· |
| AA | 2164 | hCoV-19/Denmark/DCGC-18535/2020\|EPI_ISL_750026\|202 | ···················· | ···················· | ···················· | ···················· | ···················· | ···················· |
| AA | 2165 | hCoV-19/Denmark/DCGC-18536/2020\|EPI_ISL_750027\|202 | ···················· | ···················· | ···················· | ···················· | ···················· | ···················· |
| AA | 2166 | hCoV-19/Denmark/DCGC-18538/2020\|EPI_ISL_750028\|202 | ···················· | ···················· | ···················· | ···················· | ···················· | ···················· |
| AA | 2167 | hCoV-19/Denmark/DCGC-18539/2020\|EPI_ISL_750029\|202 | ···················· | ···················· | ···················· | ···················· | ···················· | ···················· |
| AA | 2168 | hCoV-19/Denmark/DCGC-18540/2020\|EPI_ISL_750030\|202 | ···················· | ···················· | ···················· | ···················· | ···················· | ···················· |
| AA | 2169 | hCoV-19/Denmark/DCGC-18541/2020\|EPI_ISL_750031\|202 | ···················· | ···················· | ···················· | ···················· | ···················· | ···················· |
| AA | 2170 | hCoV-19/Denmark/DCGC-18542/2020\|EPI_ISL_750032\|202 | ···················· | ···················· | ···················· | ···················· | ···················· | ···················· |
| AA | 2171 | hCoV-19/Denmark/DCGC-18543/2020\|EPI_ISL_750033\|202 | ···················· | ···················· | ···················· | ···················· | ···················· | ···················· |
| AA | 2172 | hCoV-19/Denmark/DCGC-18544/2020\|EPI_ISL_750034\|202 | ···················· | ···················· | ···················· | ···················· | ···················· | ···················· |
| AA | 2173 | hCoV-19/Denmark/DCGC-18548/2020\|EPI_ISL_750035\|202 | ···················· | ···················· | ···················· | ···················· | ···················· | ···················· |
| AA | 2174 | hCoV-19/Denmark/DCGC-18549/2020\|EPI_ISL_750036\|202 | ···················· | ···················· | ···················· | ···················· | ···················· | ···················· |
| AA | 2175 | hCoV-19/Denmark/DCGC-17261/2020\|EPI_ISL_750862\|202 | ···················· | ···················· | ···················· | ···················· | ···················· | ···················· |
| AA | 2176 | hCoV-19/Denmark/DCGC-17263/2020\|EPI_ISL_750863\|202 | ···················· | ···················· | ···················· | ···················· | ···················· | ···················· |
| AA | 2177 | hCoV-19/Denmark/DCGC-17264/2020\|EPI_ISL_750864\|202 | ···················· | ···················· | ···················· | ···················· | ···················· | ···················· |
| AA | 2178 | hCoV-19/Denmark/DCGC-17265/2020\|EPI_ISL_750865\|202 | ···················· | ···················· | ···················· | ···················· | ···················· | ···················· |
| AA | 2179 | hCoV-19/Denmark/DCGC-17267/2020\|EPI_ISL_750866\|202 | ···················· | ···················· | ···················· | ···················· | ···················· | ···················· |
| AA | 2180 | hCoV-19/Denmark/DCGC-17268/2020\|EPI_ISL_750867\|202 | ···················· | ···················· | ···················· | ···················· | ···················· | ···················· |
| AA | 2181 | hCoV-19/Denmark/DCGC-17269/2020\|EPI_ISL_750868\|202 | ···················· | ···················· | ···················· | ···················· | ···················· | ···················· |
| AA | 2182 | hCoV-19/Denmark/DCGC-17270/2020\|EPI_ISL_750869\|202 | ···················· | ···················· | ···················· | ···················· | ···················· | ···················· |
| AA | 2183 | hCoV-19/Denmark/DCGC-17271/2020\|EPI_ISL_750870\|202 | ···················· | ···················· | ···················· | ···················· | ···················· | ···················· |
| AA | 2184 | hCoV-19/Denmark/DCGC-17274/2020\|EPI_ISL_750871\|202 | ···················· | ···················· | ···················· | ···················· | ···················· | ···················· |
| AA | 2185 | hCoV-19/Denmark/DCGC-17275/2020\|EPI_ISL_750872\|202 | ···················· | ···················· | ···················· | ···················· | ···················· | ···················· |
| AA | 2186 | hCoV-19/Denmark/DCGC-17276/2020\|EPI_ISL_750873\|202 | ···················· | ···················· | ···················· | ···················· | ···················· | ···················· |
| AA | 2187 | hCoV-19/Denmark/DCGC-17277/2020\|EPI_ISL_750874\|202 | ···················· | ···················· | ···················· | ···················· | ···················· | ···················· |
| AA | 2188 | hCoV-19/Denmark/DCGC-17278/2020\|EPI_ISL_750875\|202 | ···················· | ···················· | ···················· | ···················· | ···················· | ···················· |
| AA | 2189 | hCoV-19/Denmark/DCGC-17280/2020\|EPI_ISL_750876\|202 | ···················· | ···················· | ···················· | ···················· | ···················· | ···················· |
| AA | 2190 | hCoV-19/Denmark/DCGC-17281/2020\|EPI_ISL_750877\|202 | ···················· | ···················· | ···················· | ···················· | ···················· | ···················· |
| AA | 2191 | hCoV-19/Denmark/DCGC-17283/2020\|EPI_ISL_750878\|202 | ···················· | ···················· | ···················· | ···················· | ···················· | ···················· |
| AA | 2192 | hCoV-19/Denmark/DCGC-17285/2020\|EPI_ISL_750879\|202 | ···················· | ···················· | ···················· | ···················· | ···················· | ···················· |
| AA | 2193 | hCoV-19/Denmark/DCGC-17287/2020\|EPI_ISL_750880\|202 | ···················· | ···················· | ···················· | ···················· | ···················· | ···················· |
| AA | 2194 | hCoV-19/Denmark/DCGC-17289/2020\|EPI_ISL_750881\|202 | ···················· | ···················· | ···················· | ···················· | ···················· | ···················· |
| AA | 2195 | hCoV-19/Denmark/DCGC-17290/2020\|EPI_ISL_750882\|202 | ···················· | ···················· | ···················· | ···················· | ···················· | ···················· |
| AA | 2196 | hCoV-19/Denmark/DCGC-17291/2020\|EPI_ISL_750883\|202 | ···················· | ···················· | ···················· | ···················· | ···················· | ···················· |
| AA | 2197 | hCoV-19/Denmark/DCGC-17292/2020\|EPI_ISL_750884\|202 | ···················· | ···················· | ···················· | ···················· | ···················· | ···················· |
| AA | 2198 | hCoV-19/Denmark/DCGC-17296/2020\|EPI_ISL_750885\|202 | ···················· | ···················· | ···················· | ···················· | ···················· | ···················· |
| AA | 2199 | hCoV-19/Denmark/DCGC-17297/2020\|EPI_ISL_750886\|202 | ···················· | ···················· | ···················· | ···················· | ···················· | ···················· |
| AA | 2200 | hCoV-19/Denmark/DCGC-17298/2020\|EPI_ISL_750887\|202 | ···················· | ···················· | ···················· | ···················· | ···················· | ···················· |
| AA | 2201 | hCoV-19/Denmark/DCGC-17299/2020\|EPI_ISL_750888\|202 | ···················· | ···················· | ···················· | ···················· | ···················· | ···················· |
| AA | 2202 | hCoV-19/Denmark/DCGC-17300/2020\|EPI_ISL_750889\|202 | ···················· | ···················· | ···················· | ···················· | ···················· | ···················· |
| AA | 2203 | hCoV-19/Denmark/DCGC-17301/2020\|EPI_ISL_750890\|202 | ···················· | ···················· | ···················· | ···················· | ···················· | ···················· |
| AA | 2204 | hCoV-19/Denmark/DCGC-17302/2020\|EPI_ISL_750891\|202 | ···················· | ···················· | ···················· | ···················· | ···················· | ···················· |
| AA | 2205 | hCoV-19/Denmark/DCGC-17303/2020\|EPI_ISL_750892\|202 | ···················· | ···················· | ···················· | ···················· | ···················· | ···················· |
| AA | 2206 | hCoV-19/Denmark/DCGC-17304/2020\|EPI_ISL_750893\|202 | ···················· | ···················· | ···················· | ···················· | ···················· | ···················· |
| AA | 2207 | hCoV-19/Denmark/DCGC-17305/2020\|EPI_ISL_750894\|202 | ···················· | ···················· | ···················· | ···················· | ···················· | ···················· |
| AA | 2208 | hCoV-19/Denmark/DCGC-17306/2020\|EPI_ISL_750895\|202 | ···················· | ···················· | ···················· | ···················· | ···················· | ···················· |
| AA | 2209 | hCoV-19/Denmark/DCGC-17308/2020\|EPI_ISL_750896\|202 | ···················· | ···················· | ···················· | ···················· | ···················· | ···················· |
| AA | 2210 | hCoV-19/Denmark/DCGC-17312/2020\|EPI_ISL_750897\|202 | ···················· | ···················· | ···················· | ···················· | ···················· | ···················· |
| AA | 2211 | hCoV-19/Denmark/DCGC-17313/2020\|EPI_ISL_750898\|202 | ···················· | ···················· | ···················· | ···················· | ···················· | ···················· |
| AA | 2212 | hCoV-19/Denmark/DCGC-17314/2020\|EPI_ISL_750899\|202 | ···················· | ···················· | ···················· | ···················· | ···················· | ···················· |
| AA | 2213 | hCoV-19/Italy/VEN-UniVR-47/2020\|EPI_ISL_751357\|202 | ···················· | ···················· | ···················· | ···················· | ···················· | ···················· |
| AA | 2214 | hCoV-19/Italy/VEN-UniVR-48/2020\|EPI_ISL_751358\|202 | ···················· | ···················· | ···················· | ···················· | ···················· | ···················· |
| AA | 2215 | hCoV-19/Italy/VEN-UniVR-49/2020\|EPI_ISL_751359\|202 | ···················· | ···················· | ···················· | ···················· | ···················· | ···················· |
| AA | 2216 | hCoV-19/Italy/VEN-UniVR-50/2020\|EPI_ISL_751360\|202 | ···················· | ···················· | ···················· | ···················· | ···················· | ···················· |
| AA | 2217 | hCoV-19/Italy/VEN-UniVR-51/2020\|EPI_ISL_751361\|202 | ···················· | ···················· | ···················· | ···················· | ···················· | ···················· |
| AA | 2218 | hCoV-19/Italy/VEN-UniVR-52/2020\|EPI_ISL_751362\|202 | ···················· | ···················· | ···················· | ···················· | ···················· | ···················· |
| AA | 2219 | hCoV-19/Italy/VEN-UniVR-53/2020\|EPI_ISL_751363\|202 | ···················· | ···················· | ···················· | ···················· | ···················· | ···················· |
| AA | 2220 | hCoV-19/Italy/VEN-UniVR-54/2020\|EPI_ISL_751364\|202 | ···················· | ···················· | ···················· | ···················· | ···················· | ···················· |
| AA | 2221 | hCoV-19/Italy/VEN-UniVR-55/2020\|EPI_ISL_751365\|202 | ···················· | ···················· | ···················· | ···················· | ···················· | ···················· |
| AA | 2222 | hCoV-19/Italy/VEN-UniVR-56/2020\|EPI_ISL_751366\|202 | ···················· | ···················· | ···················· | ···················· | ···················· | ···················· |
| AA | 2223 | hCoV-19/Italy/VEN-UniVR-57/2020\|EPI_ISL_751367\|202 | ···················· | ···················· | ···················· | ···················· | ···················· | ···················· |
| AA | 2224 | hCoV-19/USA/HI-H200125/2020\|EPI_ISL_752688\|2020-06 | ···················· | ···················· | ···················· | ···················· | ···················· | ···················· |
| AA | 2225 | hCoV-19/USA/HI-H200126/2020\|EPI_ISL_752689\|2020-06 | ···················· | ···················· | ···················· | ···················· | ···················· | ···················· |
| AA | 2226 | hCoV-19/USA/HI-H200127/2020\|EPI_ISL_752690\|2020-06 | ···················· | ···················· | ···················· | ···················· | ···················· | ···················· |
| AA | 2227 | hCoV-19/USA/HI-H200128/2020\|EPI_ISL_752691\|2020-06 | ···················· | ···················· | ···················· | ···················· | ···················· | ···················· |
| AA | 2228 | hCoV-19/USA/HI-H200129/2020\|EPI_ISL_752692\|2020-07 | ···················· | ···················· | ···················· | ···················· | ···················· | ···················· |
| AA | 2229 | hCoV-19/USA/HI-H200130/2020\|EPI_ISL_752693\|2020-07 | ···················· | ···················· | ···················· | ···················· | ···················· | ···················· |
| AA | 2230 | hCoV-19/USA/HI-H200131/2020\|EPI_ISL_752694\|2020-07 | ···················· | ···················· | ···················· | ···················· | ···················· | ···················· |
| AA | 2231 | hCoV-19/USA/HI-H200132/2020\|EPI_ISL_752695\|2020-07 | ···················· | ···················· | ···················· | ···················· | ···················· | ···················· |
| AA | 2232 | hCoV-19/USA/HI-H200133/2020\|EPI_ISL_752696\|2020-07 | ···················· | ·A·················· | ···················· | ···················· | ···················· | ···················· |
| AA | 2233 | hCoV-19/USA/HI-H200134/2020\|EPI_ISL_752697\|2020-07 | ···················· | ···················· | ···················· | ···················· | ···················· | ···················· |
| AA | 2234 | hCoV-19/USA/HI-H200135/2020\|EPI_ISL_752698\|2020-07 | ···················· | ···················· | ···················· | ···················· | ···················· | ···················· |
| AA | 2235 | hCoV-19/Denmark/DCGC-20518/2020\|EPI_ISL_759200\|202 | ···················· | ···················· | ···················· | ···················· | ···················· | ···············A···· |
| AA | 2236 | hCoV-19/Denmark/DCGC-20523/2020\|EPI_ISL_759201\|202 | ···················· | ···················· | ···················· | ···················· | ···················· | ···················· |
| AA | 2237 | hCoV-19/Denmark/DCGC-20525/2020\|EPI_ISL_759202\|202 | ···················· | ···················· | ···················· | ···················· | ···················· | ···················· |
| AA | 2238 | hCoV-19/Denmark/DCGC-20526/2020\|EPI_ISL_759203\|202 | ···················· | ···················· | ···················· | ···················· | ···················· | ···················· |
| AA | 2239 | hCoV-19/Denmark/DCGC-20529/2020\|EPI_ISL_759204\|202 | ···················· | ···················· | ···················· | ···················· | ···················· | ···················· |
| AA | 2240 | hCoV-19/Denmark/DCGC-20530/2020\|EPI_ISL_759205\|202 | ···················· | ···················· | ···················· | ···················· | ···················· | ···················· |
| AA | 2241 | hCoV-19/Denmark/DCGC-20532/2020\|EPI_ISL_759206\|202 | ···················· | ···················· | ···················· | ···················· | ···················· | ···················· |
| AA | 2242 | hCoV-19/Denmark/DCGC-20533/2020\|EPI_ISL_759207\|202 | ···················· | ···················· | ···················· | ···················· | ···················· | ···················· |

| | | | | | | | |
|---|---|---|---|---|---|---|---|
| AA | 2243 hCoV-19/Denmark/DCGC-20535/2020\|EPI_ISL_759208\|202 | ···················· | ···················· | ···················· | ···················· | ···················· | ···················· |
| AA | 2244 hCoV-19/Denmark/DCGC-20538/2020\|EPI_ISL_759209\|202 | ···················· | ···················· | ···················· | ···················· | ···················· | ···················· |
| AA | 2245 hCoV-19/Denmark/DCGC-20541/2020\|EPI_ISL_759210\|202 | ···················· | ···················· | ···················· | ···················· | ···················· | ···················· |
| AA | 2246 hCoV-19/Denmark/DCGC-20547/2020\|EPI_ISL_759211\|202 | ···················· | ···················· | ···················· | ···················· | ···················· | ···················· |
| AA | 2247 hCoV-19/Denmark/DCGC-20549/2020\|EPI_ISL_759212\|202 | ···················· | ···················· | ···················· | ···················· | ···················· | ···················· |
| AA | 2248 hCoV-19/Denmark/DCGC-20551/2020\|EPI_ISL_759213\|202 | ···················· | ···················· | ···················· | ···················· | ···················· | ···················· |
| AA | 2249 hCoV-19/Denmark/DCGC-20552/2020\|EPI_ISL_759214\|202 | ···················· | ···················· | ···················· | ···················· | ···················· | ···················· |
| AA | 2250 hCoV-19/Denmark/DCGC-20553/2020\|EPI_ISL_759215\|202 | ···················· | ···················· | ···················· | ···················· | ···················· | ···················· |
| AA | 2251 hCoV-19/Denmark/DCGC-20554/2020\|EPI_ISL_759216\|202 | ···················· | ···················· | ···················· | ···················· | ···················· | ···················· |
| AA | 2252 hCoV-19/Denmark/DCGC-20555/2020\|EPI_ISL_759217\|202 | ···················· | ···················· | ···················· | ···················· | ···················· | ···················· |
| AA | 2253 hCoV-19/Denmark/DCGC-20557/2020\|EPI_ISL_759218\|202 | ···················· | ···················· | ···················· | ···················· | ···················· | ···················· |
| AA | 2254 hCoV-19/Denmark/DCGC-20558/2020\|EPI_ISL_759219\|202 | ···················· | ···················· | ···················· | ···················· | ···················· | ···················· |
| AA | 2255 hCoV-19/Denmark/DCGC-20561/2020\|EPI_ISL_759220\|202 | ···················· | ···················· | ···················· | ···················· | ···················· | ···················· |
| AA | 2256 hCoV-19/Denmark/DCGC-20562/2020\|EPI_ISL_759221\|202 | ···················· | ···················· | ···················· | ···················· | ···················· | ···················· |
| AA | 2257 hCoV-19/Denmark/DCGC-20565/2020\|EPI_ISL_759222\|202 | ···················· | ···················· | ···················· | ···················· | ···················· | ···················· |
| AA | 2258 hCoV-19/Denmark/DCGC-20568/2020\|EPI_ISL_759223\|202 | ···················· | ···················· | ···················· | ···················· | ···················· | ···················· |
| AA | 2259 hCoV-19/Denmark/DCGC-20569/2020\|EPI_ISL_759224\|202 | ···················· | ···················· | ···················· | ···················· | ···················· | ···················· |
| AA | 2260 hCoV-19/Denmark/DCGC-20570/2020\|EPI_ISL_759225\|202 | ···················· | ···················· | ···················· | ···················· | ···················· | ···················· |
| AA | 2261 hCoV-19/Denmark/DCGC-20572/2020\|EPI_ISL_759226\|202 | ···················· | ···················· | ···················· | ···················· | ···················· | ···················· |
| AA | 2262 hCoV-19/Denmark/DCGC-20573/2020\|EPI_ISL_759227\|202 | ···················· | ···················· | ···················· | ···················· | ···················· | ···················· |
| AA | 2263 hCoV-19/Denmark/DCGC-20574/2020\|EPI_ISL_759228\|202 | ···················· | ···················· | ···················· | ···················· | ···················· | ···················· |
| AA | 2264 hCoV-19/Denmark/DCGC-20575/2020\|EPI_ISL_759229\|202 | ···················· | ···················· | ···················· | ···················· | ···················· | ···················· |
| AA | 2265 hCoV-19/Denmark/DCGC-20576/2020\|EPI_ISL_759230\|202 | ···················· | ···················· | ···················· | ···················· | ···················· | ···················· |
| AA | 2266 hCoV-19/Denmark/DCGC-20578/2020\|EPI_ISL_759231\|202 | ···················· | ···················· | ···················· | ···················· | ···················· | ···················· |
| AA | 2267 hCoV-19/Denmark/DCGC-20579/2020\|EPI_ISL_759232\|202 | ···················· | ···················· | ···················· | ···················· | ···················· | ···················· |
| AA | 2268 hCoV-19/Denmark/DCGC-20580/2020\|EPI_ISL_759233\|202 | ···················· | ···················· | ···················· | ···················· | ···················· | ···················· |
| AA | 2269 hCoV-19/Denmark/DCGC-20581/2020\|EPI_ISL_759234\|202 | ···················· | ···················· | ···················· | ···················· | ···················· | ···················· |
| AA | 2270 hCoV-19/Denmark/DCGC-20582/2020\|EPI_ISL_759235\|202 | ···················· | ···················· | ···················· | ···················· | ···················· | ···················· |
| AA | 2271 hCoV-19/Denmark/DCGC-20584/2020\|EPI_ISL_759236\|202 | ···················· | ···················· | ···················· | ···················· | ···················· | ···················· |
| AA | 2272 hCoV-19/Denmark/DCGC-20585/2020\|EPI_ISL_759237\|202 | ···················· | ···················· | ···················· | ···················· | ···················· | ···················· |
| AA | 2273 hCoV-19/Denmark/DCGC-20586/2020\|EPI_ISL_759238\|202 | ···················· | ···················· | ···················· | ···················· | ···················· | ···················· |
| AA | 2274 hCoV-19/Denmark/DCGC-20589/2020\|EPI_ISL_759239\|202 | ···················· | ···················· | ···················· | ···················· | ···················· | ···················· |
| AA | 2275 hCoV-19/Denmark/DCGC-20605/2020\|EPI_ISL_759250\|202 | ···················· | ···················· | ···················· | ···················· | ···················· | ···················· |
| AA | 2276 hCoV-19/Denmark/DCGC-20610/2020\|EPI_ISL_759251\|202 | ···················· | ···················· | ···················· | ···················· | ···················· | ···················· |
| AA | 2277 hCoV-19/Denmark/DCGC-20611/2020\|EPI_ISL_759252\|202 | ···················· | ···················· | ···················· | ···················· | ···················· | ···················· |
| AA | 2278 hCoV-19/Denmark/DCGC-20612/2020\|EPI_ISL_759253\|202 | ···················· | ···················· | ···················· | ···················· | ···················· | ···················· |
| AA | 2279 hCoV-19/Denmark/DCGC-20614/2020\|EPI_ISL_759254\|202 | ···················· | ···················· | ···················· | ···················· | ···················· | ···················· |
| AA | 2280 hCoV-19/Denmark/DCGC-20615/2020\|EPI_ISL_759255\|202 | ···················· | ···················· | ···················· | ···················· | ···················· | ···················· |
| AA | 2281 hCoV-19/Denmark/DCGC-20616/2020\|EPI_ISL_759256\|202 | ···················· | ···················· | ···················· | ···················· | ···················· | ···················· |
| AA | 2282 hCoV-19/Denmark/DCGC-20617/2020\|EPI_ISL_759257\|202 | ···················· | ···················· | ···················· | ···················· | ···················· | ···················· |
| AA | 2283 hCoV-19/Denmark/DCGC-20619/2020\|EPI_ISL_759258\|202 | ···················· | ···················· | ···················· | ···················· | ···················· | ···················· |
| AA | 2284 hCoV-19/Denmark/DCGC-20622/2020\|EPI_ISL_759259\|202 | ···················· | ···················· | ···················· | ···················· | ···················· | ···················· |
| AA | 2285 hCoV-19/Denmark/DCGC-20623/2020\|EPI_ISL_759260\|202 | ···················· | ···················· | ···················· | ···················· | ···················· | ···················· |
| AA | 2286 hCoV-19/Denmark/DCGC-20624/2020\|EPI_ISL_759261\|202 | ···················· | ···················· | ···················· | ···················· | ···················· | ···················· |
| AA | 2287 hCoV-19/Denmark/DCGC-20625/2020\|EPI_ISL_759262\|202 | ···················· | ···················· | ···················· | ···················· | ···················· | ···················· |
| AA | 2288 hCoV-19/Denmark/DCGC-20626/2020\|EPI_ISL_759263\|202 | ···················· | ···················· | ···················· | ···················· | ···················· | ···················· |
| AA | 2289 hCoV-19/Denmark/DCGC-20627/2020\|EPI_ISL_759264\|202 | ···················· | ···················· | ···················· | ···················· | ···················· | ···················· |
| AA | 2290 hCoV-19/Denmark/DCGC-20628/2020\|EPI_ISL_759265\|202 | ···················· | ···················· | ···················· | ···················· | ···················· | ···················· |
| AA | 2291 hCoV-19/Denmark/DCGC-20637/2020\|EPI_ISL_759266\|202 | ···················· | ···················· | ···················· | ···················· | ···················· | ···················· |
| AA | 2292 hCoV-19/Denmark/DCGC-20638/2020\|EPI_ISL_759267\|202 | ···················· | ···················· | ···················· | ···················· | ···················· | ···················· |
| AA | 2293 hCoV-19/Denmark/DCGC-20639/2020\|EPI_ISL_759268\|202 | ···················· | ·A·················· | ···················· | ···················· | ···················· | ···A················ |
| AA | 2294 hCoV-19/Denmark/DCGC-20640/2020\|EPI_ISL_759269\|202 | ···················· | ···················· | ···················· | ···················· | ···················· | ···················· |
| AA | 2295 hCoV-19/Denmark/DCGC-20641/2020\|EPI_ISL_759270\|202 | ···················· | ···················· | ···················· | ···················· | ···················· | ···················· |
| AA | 2296 hCoV-19/Denmark/DCGC-20642/2020\|EPI_ISL_759271\|202 | ···················· | ···················· | ···················· | ···················· | ···················· | ···················· |
| AA | 2297 hCoV-19/Denmark/DCGC-20644/2020\|EPI_ISL_759272\|202 | ···················· | ···················· | ···················· | ···················· | ···················· | ···················· |
| AA | 2298 hCoV-19/Denmark/DCGC-20645/2020\|EPI_ISL_759273\|202 | ···················· | ···················· | ···················· | ···················· | ···················· | ···················· |
| AA | 2299 hCoV-19/Denmark/DCGC-20648/2020\|EPI_ISL_759274\|202 | ···················· | ···················· | ···················· | ···················· | ···················· | ···················· |
| AA | 2300 hCoV-19/Denmark/DCGC-20650/2020\|EPI_ISL_759275\|202 | ···················· | ···················· | ···················· | ···················· | ···················· | ···················· |
| AA | 2301 hCoV-19/Denmark/DCGC-20652/2020\|EPI_ISL_759276\|202 | ···················· | ···················· | ···················· | ···················· | ···················· | ···················· |
| AA | 2302 hCoV-19/Denmark/DCGC-20653/2020\|EPI_ISL_759277\|202 | ···················· | ···················· | ···················· | ···················· | ···················· | ···················· |
| AA | 2303 hCoV-19/Denmark/DCGC-20654/2020\|EPI_ISL_759278\|202 | ···················· | ···················· | ···················· | ···················· | ···················· | ···················· |
| AA | 2304 hCoV-19/Denmark/DCGC-20656/2020\|EPI_ISL_759279\|202 | ···················· | ···················· | ···················· | ···················· | ···················· | ···················· |
| AA | 2305 hCoV-19/Denmark/DCGC-19514/2020\|EPI_ISL_749000\|202 | ···················· | ···················· | ···················· | ···················· | ···················· | ···················· |
| AA | 2306 hCoV-19/Denmark/DCGC-19515/2020\|EPI_ISL_749001\|202 | ···················· | ···················· | ···················· | ···················· | ···················· | ···················· |
| AA | 2307 hCoV-19/Denmark/DCGC-19516/2020\|EPI_ISL_749002\|202 | ···················· | ···················· | ···················· | ···················· | ···················· | ···················· |
| AA | 2308 hCoV-19/Denmark/DCGC-19517/2020\|EPI_ISL_749003\|202 | ···················· | ···················· | ···················· | ···················· | ···················· | ···················· |
| AA | 2309 hCoV-19/Denmark/DCGC-19518/2020\|EPI_ISL_749004\|202 | ···················· | ···················· | ···················· | ···················· | ···················· | ···················· |
| AA | 2310 hCoV-19/Denmark/DCGC-19519/2020\|EPI_ISL_749005\|202 | ···················· | ···················· | ···················· | ···················· | ···················· | ···················· |
| AA | 2311 hCoV-19/Denmark/DCGC-19522/2020\|EPI_ISL_749006\|202 | ···················· | ···················· | ···················· | ···················· | ···················· | ···················· |
| AA | 2312 hCoV-19/Denmark/DCGC-19525/2020\|EPI_ISL_749007\|202 | ···················· | ···················· | ···················· | ···················· | ···················· | ···················· |
| AA | 2313 hCoV-19/Denmark/DCGC-19527/2020\|EPI_ISL_749008\|202 | ···················· | ···················· | ···················· | ···················· | ···················· | ···················· |
| AA | 2314 hCoV-19/Denmark/DCGC-19528/2020\|EPI_ISL_749009\|202 | ···················· | ···················· | ···················· | ···················· | ···················· | ···················· |
| AA | 2315 hCoV-19/Denmark/DCGC-19530/2020\|EPI_ISL_749010\|202 | ···················· | ···················· | ···················· | ···················· | ···················· | ···················· |
| AA | 2316 hCoV-19/Denmark/DCGC-19532/2020\|EPI_ISL_749011\|202 | ···················· | ···················· | ···················· | ···················· | ···················· | ···················· |
| AA | 2317 hCoV-19/Denmark/DCGC-19534/2020\|EPI_ISL_749012\|202 | ···················· | ···················· | ···················· | ···················· | ···················· | ···················· |
| AA | 2318 hCoV-19/Denmark/DCGC-19535/2020\|EPI_ISL_749013\|202 | ···················· | ···················· | ···················· | ···················· | ···················· | ···················· |
| AA | 2319 hCoV-19/Denmark/DCGC-19536/2020\|EPI_ISL_749014\|202 | ···················· | ···················· | ···················· | ···················· | ···················· | ···················· |
| AA | 2320 hCoV-19/Denmark/DCGC-19538/2020\|EPI_ISL_749015\|202 | ···················· | ···················· | ···················· | ···················· | ···················· | ···················· |
| AA | 2321 hCoV-19/Denmark/DCGC-19542/2020\|EPI_ISL_749016\|202 | ···················· | ···················· | ···················· | ···················· | ···················· | ···················· |
| AA | 2322 hCoV-19/Denmark/DCGC-19543/2020\|EPI_ISL_749017\|202 | ···················· | ···················· | ···················· | ···················· | ···················· | ···················· |

| | | | | | | | |
|---|---|---|---|---|---|---|---|
| AA | 2323 hCoV-19/Denmark/DCGC-19547/2020\|EPI_ISL_749018\|202 | .................... | .................... | ..................... | .................... | .................... | .................... |
| AA | 2324 hCoV-19/Denmark/DCGC-19550/2020\|EPI_ISL_749019\|202 | .................... | .................... | ..................... | .................... | .................... | .................... |
| AA | 2325 hCoV-19/Denmark/DCGC-19551/2020\|EPI_ISL_749020\|202 | .................... | .................... | ..................... | .................... | .................... | .................... |
| AA | 2326 hCoV-19/Denmark/DCGC-19553/2020\|EPI_ISL_749021\|202 | .................... | .................... | ..................... | .................... | .................... | .................... |
| AA | 2327 hCoV-19/Denmark/DCGC-19554/2020\|EPI_ISL_749022\|202 | .................... | .................... | ..................... | .................... | .................... | .................... |
| AA | 2328 hCoV-19/Denmark/DCGC-19555/2020\|EPI_ISL_749023\|202 | .................... | .................... | ..................... | .................... | .................... | .................... |
| AA | 2329 hCoV-19/Denmark/DCGC-19556/2020\|EPI_ISL_749024\|202 | .................... | .................... | ..................... | .................... | .................... | .................... |
| AA | 2330 hCoV-19/Denmark/DCGC-19557/2020\|EPI_ISL_749025\|202 | .................... | .................... | ..................... | .................... | .................... | .................... |
| AA | 2331 hCoV-19/Denmark/DCGC-19561/2020\|EPI_ISL_749026\|202 | .................... | .................... | ..................... | .................... | .................... | .................... |
| AA | 2332 hCoV-19/Denmark/DCGC-19562/2020\|EPI_ISL_749027\|202 | .................... | .................... | ..................... | .................... | .................... | .................... |
| AA | 2333 hCoV-19/Denmark/DCGC-19563/2020\|EPI_ISL_749028\|202 | .................... | .................... | ..................... | .................... | .................... | .................... |
| AA | 2334 hCoV-19/Denmark/DCGC-19564/2020\|EPI_ISL_749029\|202 | .................... | .................... | ..................... | .................... | .................... | .................... |
| AA | 2335 hCoV-19/Denmark/DCGC-19581/2020\|EPI_ISL_749040\|202 | .................... | .................... | ..................... | .................... | .................... | .................... |
| AA | 2336 hCoV-19/Denmark/DCGC-19582/2020\|EPI_ISL_749041\|202 | .................... | .................... | ..................... | .................... | .................... | .................... |
| AA | 2337 hCoV-19/Denmark/DCGC-19583/2020\|EPI_ISL_749042\|202 | .................... | .A.................. | ..................... | .................... | .................... | .................... |
| AA | 2338 hCoV-19/Denmark/DCGC-19585/2020\|EPI_ISL_749043\|202 | .................... | .................... | ..................... | .................... | .................... | .................... |
| AA | 2339 hCoV-19/Denmark/DCGC-19586/2020\|EPI_ISL_749044\|202 | .................... | .................... | ..................... | .................... | .................... | .................... |
| AA | 2340 hCoV-19/Denmark/DCGC-19587/2020\|EPI_ISL_749045\|202 | .................... | .................... | ..................... | .................... | .................... | .................... |
| AA | 2341 hCoV-19/Denmark/DCGC-19590/2020\|EPI_ISL_749046\|202 | .................... | .................... | ..................... | .................... | .................... | .................... |
| AA | 2342 hCoV-19/Denmark/DCGC-19594/2020\|EPI_ISL_749047\|202 | .................... | .................... | ..................... | .................... | .................... | .................... |
| AA | 2343 hCoV-19/Denmark/DCGC-19595/2020\|EPI_ISL_749048\|202 | .................... | .................... | ..................... | .................... | .................... | .................... |
| AA | 2344 hCoV-19/Denmark/DCGC-19596/2020\|EPI_ISL_749049\|202 | .................... | .................... | ..................... | .................... | .................... | .................... |
| AA | 2345 hCoV-19/Denmark/DCGC-19597/2020\|EPI_ISL_749050\|202 | .................... | .................... | ..................... | .................... | .................... | .................... |
| AA | 2346 hCoV-19/Denmark/DCGC-19599/2020\|EPI_ISL_749051\|202 | .................... | .................... | ..................... | .................... | .................... | .................... |
| AA | 2347 hCoV-19/Denmark/DCGC-19600/2020\|EPI_ISL_749052\|202 | .................... | .................... | ..................... | .................... | .................... | .................... |
| AA | 2348 hCoV-19/Denmark/DCGC-19602/2020\|EPI_ISL_749053\|202 | .................... | .................... | ..................... | .................... | .................... | .................... |
| AA | 2349 hCoV-19/Denmark/DCGC-19604/2020\|EPI_ISL_749054\|202 | .................... | .................... | ..................... | .................... | .................... | .................... |
| AA | 2350 hCoV-19/Denmark/DCGC-19606/2020\|EPI_ISL_749055\|202 | .................... | .................... | ..................... | .................... | .................... | .................... |
| AA | 2351 hCoV-19/Denmark/DCGC-19607/2020\|EPI_ISL_749056\|202 | .................... | .................... | ..................... | .................... | .................... | .................... |
| AA | 2352 hCoV-19/Denmark/DCGC-19609/2020\|EPI_ISL_749057\|202 | .................... | .................... | ..................... | .................... | .................... | .................... |
| AA | 2353 hCoV-19/Denmark/DCGC-19610/2020\|EPI_ISL_749058\|202 | .................... | .................... | ..................... | .................... | .................... | .................... |
| AA | 2354 hCoV-19/Denmark/DCGC-19611/2020\|EPI_ISL_749059\|202 | .................... | .................... | ..................... | .................... | .................... | .................... |
| AA | 2355 hCoV-19/Denmark/DCGC-19612/2020\|EPI_ISL_749060\|202 | .................... | .................... | ..................... | .................... | .................... | .................... |
| AA | 2356 hCoV-19/Denmark/DCGC-19613/2020\|EPI_ISL_749061\|202 | .................... | .................... | ..................... | .................... | .................... | .................... |
| AA | 2357 hCoV-19/Denmark/DCGC-19614/2020\|EPI_ISL_749062\|202 | .................... | .................... | ..................... | .................... | .................... | .................... |
| AA | 2358 hCoV-19/Denmark/DCGC-19616/2020\|EPI_ISL_749063\|202 | .................... | .................... | ..................... | .................... | .................... | .................... |
| AA | 2359 hCoV-19/Denmark/DCGC-19619/2020\|EPI_ISL_749064\|202 | .................... | .................... | ..................... | .................... | .................... | .................... |
| AA | 2360 hCoV-19/Denmark/DCGC-19622/2020\|EPI_ISL_749065\|202 | .................... | .................... | ..................... | .................... | .................... | .................... |
| AA | 2361 hCoV-19/Denmark/DCGC-19624/2020\|EPI_ISL_749066\|202 | .................... | .................... | ..................... | .................... | .................... | .................... |
| AA | 2362 hCoV-19/Denmark/DCGC-19625/2020\|EPI_ISL_749067\|202 | .................... | .................... | ..................... | .................... | .................... | .................... |
| AA | 2363 hCoV-19/Denmark/DCGC-19627/2020\|EPI_ISL_749068\|202 | .................... | .................... | ..................... | .................... | .................... | .................... |
| AA | 2364 hCoV-19/Denmark/DCGC-19628/2020\|EPI_ISL_749069\|202 | .................... | .................... | ..................... | .................... | .................... | .................... |
| AA | 2365 hCoV-19/Denmark/DCGC-19630/2020\|EPI_ISL_749070\|202 | .................... | .................... | ..................... | .................... | .................... | .................... |
| AA | 2366 hCoV-19/Denmark/DCGC-19631/2020\|EPI_ISL_749071\|202 | .................... | .................... | ..................... | .................... | .................... | .................... |
| AA | 2367 hCoV-19/Denmark/DCGC-19632/2020\|EPI_ISL_749072\|202 | .................... | .................... | ..................... | .................... | .................... | .................... |
| AA | 2368 hCoV-19/Denmark/DCGC-19633/2020\|EPI_ISL_749073\|202 | .................... | .................... | ..................... | .................... | .................... | .................... |
| AA | 2369 hCoV-19/Denmark/DCGC-19636/2020\|EPI_ISL_749074\|202 | .................... | .................... | ..................... | .................... | .................... | .................... |
| AA | 2370 hCoV-19/Denmark/DCGC-19637/2020\|EPI_ISL_749075\|202 | .................... | .A.................. | ..................... | .................... | .................... | .................... |
| AA | 2371 hCoV-19/Denmark/DCGC-19638/2020\|EPI_ISL_749076\|202 | .................... | .................... | ..................... | .................... | .................... | .................... |
| AA | 2372 hCoV-19/Denmark/DCGC-19640/2020\|EPI_ISL_749077\|202 | .................... | .................... | ..................... | .................... | .................... | .................... |
| AA | 2373 hCoV-19/Denmark/DCGC-19641/2020\|EPI_ISL_749078\|202 | .................... | .................... | ..................... | .................... | .................... | .................... |
| AA | 2374 hCoV-19/Denmark/DCGC-19643/2020\|EPI_ISL_749079\|202 | .................... | .................... | ..................... | .................... | .................... | .................... |
| AA | 2375 hCoV-19/Denmark/DCGC-18493/2020\|EPI_ISL_750000\|202 | .................... | .................... | ..................... | .................... | .................... | .................... |
| AA | 2376 hCoV-19/Denmark/DCGC-18495/2020\|EPI_ISL_750001\|202 | .................... | .................... | ..................... | .................... | .................... | .................... |
| AA | 2377 hCoV-19/Denmark/DCGC-18497/2020\|EPI_ISL_750002\|202 | .................... | .................... | ..................... | .................... | .................... | .................... |
| AA | 2378 hCoV-19/Denmark/DCGC-18500/2020\|EPI_ISL_750003\|202 | .................... | .................... | ..................... | .................... | .................... | .................... |
| AA | 2379 hCoV-19/Denmark/DCGC-18501/2020\|EPI_ISL_750004\|202 | .................... | .................... | ..................... | .................... | .................... | .................... |
| AA | 2380 hCoV-19/Denmark/DCGC-18502/2020\|EPI_ISL_750005\|202 | .................... | .................... | ..................... | .................... | .................... | .................... |
| AA | 2381 hCoV-19/Denmark/DCGC-18503/2020\|EPI_ISL_750006\|202 | .................... | .................... | ..................... | .................... | .................... | .................... |
| AA | 2382 hCoV-19/Denmark/DCGC-18504/2020\|EPI_ISL_750007\|202 | .................... | .................... | ..................... | .................... | .................... | .................... |
| AA | 2383 hCoV-19/Denmark/DCGC-18505/2020\|EPI_ISL_750008\|202 | .................... | .................... | ..................... | .................... | .................... | .................... |
| AA | 2384 hCoV-19/Denmark/DCGC-18506/2020\|EPI_ISL_750009\|202 | .................... | .................... | ..................... | .................... | .................... | .................... |
| AA | 2385 hCoV-19/Denmark/DCGC-18508/2020\|EPI_ISL_750010\|202 | .................... | .................... | ..................... | .................... | .................... | .................... |
| AA | 2386 hCoV-19/Denmark/DCGC-18510/2020\|EPI_ISL_750011\|202 | .................... | .................... | ..................... | .................... | .................... | .................... |
| AA | 2387 hCoV-19/Denmark/DCGC-18512/2020\|EPI_ISL_750012\|202 | .................... | .................... | ..................... | .................... | .................... | .................... |
| AA | 2388 hCoV-19/Denmark/DCGC-18513/2020\|EPI_ISL_750013\|202 | .................... | .................... | ..................... | .................... | .................... | .................... |
| AA | 2389 hCoV-19/Denmark/DCGC-18514/2020\|EPI_ISL_750014\|202 | .................... | .................... | ..................... | .................... | .................... | .................... |
| AA | 2390 hCoV-19/Denmark/DCGC-18516/2020\|EPI_ISL_750015\|202 | .................... | .................... | ..................... | .................... | .................... | .................... |
| AA | 2391 hCoV-19/Denmark/DCGC-18517/2020\|EPI_ISL_750016\|202 | .................... | .................... | ..................... | .................... | .................... | .................... |
| AA | 2392 hCoV-19/Denmark/DCGC-18518/2020\|EPI_ISL_750017\|202 | .................... | .................... | ..................... | .................... | .................... | .................... |
| AA | 2393 hCoV-19/Denmark/DCGC-18520/2020\|EPI_ISL_750018\|202 | .................... | .................... | ..................... | .................... | .................... | .................... |
| AA | 2394 hCoV-19/Denmark/DCGC-18523/2020\|EPI_ISL_750019\|202 | .................... | .................... | ..................... | .................... | .................... | .................... |
| AA | 2395 hCoV-19/Denmark/DCGC-18527/2020\|EPI_ISL_750022\|202 | .................... | .................... | ..................... | .................... | .................... | .................... |
| AA | 2396 hCoV-19/Denmark/DCGC-18532/2020\|EPI_ISL_750024\|202 | .................... | .................... | ..................... | .................... | .................... | .................... |
| AA | 2397 hCoV-19/Denmark/DCGC-18534/2020\|EPI_ISL_750025\|202 | .................... | .................... | ..................... | .................... | .................... | .................... |
| AA | 2398 hCoV-19/Denmark/DCGC-18550/2020\|EPI_ISL_750037\|202 | .................... | .................... | ..................... | .................... | .................... | .................... |
| AA | 2399 hCoV-19/Denmark/DCGC-18552/2020\|EPI_ISL_750038\|202 | .................... | .................... | ..................... | .................... | .................... | .................... |
| AA | 2400 hCoV-19/Denmark/DCGC-18553/2020\|EPI_ISL_750039\|202 | .................... | .................... | ..................... | .................... | .................... | .................... |
| AA | 2401 hCoV-19/Denmark/DCGC-18555/2020\|EPI_ISL_750040\|202 | .................... | .................... | ..................... | .................... | .................... | .................... |
| AA | 2402 hCoV-19/Denmark/DCGC-18556/2020\|EPI_ISL_750041\|202 | .................... | .................... | ..................... | .................... | .................... | .................... |

| | | | | | | | | |
|---|---|---|---|---|---|---|---|---|
| AA | 2403 | hCoV-19/Denmark/DCGC-18557/2020\|EPI_ISL_750042\|202 | ···················· | ···················· | ····················· | ···················· | ···················· | ···················· |
| AA | 2404 | hCoV-19/Denmark/DCGC-18559/2020\|EPI_ISL_750043\|202 | ···················· | ···················· | ····················· | ···················· | ···················· | ···················· |
| AA | 2405 | hCoV-19/Denmark/DCGC-18560/2020\|EPI_ISL_750044\|202 | ···················· | ···················· | ····················· | ···················· | ···················· | ···················· |
| AA | 2406 | hCoV-19/Denmark/DCGC-18561/2020\|EPI_ISL_750045\|202 | ···················· | ···················· | ····················· | ···················· | ···················· | ···················· |
| AA | 2407 | hCoV-19/Denmark/DCGC-18562/2020\|EPI_ISL_750046\|202 | ···················· | ···················· | ····················· | ···················· | ···················· | ···················· |
| AA | 2408 | hCoV-19/Denmark/DCGC-18563/2020\|EPI_ISL_750047\|202 | ···················· | ···················· | ····················· | ···················· | ···················· | ···················· |
| AA | 2409 | hCoV-19/Denmark/DCGC-18565/2020\|EPI_ISL_750048\|202 | ···················· | ···················· | ····················· | ···················· | ···················· | ···················· |
| AA | 2410 | hCoV-19/Denmark/DCGC-18566/2020\|EPI_ISL_750049\|202 | ···················· | ···················· | ····················· | ···················· | ···················· | ···················· |
| AA | 2411 | hCoV-19/Denmark/DCGC-18567/2020\|EPI_ISL_750050\|202 | ···················· | ···················· | ····················· | ···················· | ···················· | ···················· |
| AA | 2412 | hCoV-19/Denmark/DCGC-18570/2020\|EPI_ISL_750051\|202 | ···················· | ···················· | ····················· | ···················· | ···················· | ···················· |
| AA | 2413 | hCoV-19/Denmark/DCGC-18571/2020\|EPI_ISL_750052\|202 | ···················· | ·A·················· | ····················· | ···················· | ···················· | ··A················· |
| AA | 2414 | hCoV-19/Denmark/DCGC-18573/2020\|EPI_ISL_750053\|202 | ···················· | ···················· | ····················· | ···················· | ···················· | ··A················· |
| AA | 2415 | hCoV-19/Denmark/DCGC-18574/2020\|EPI_ISL_750054\|202 | ···················· | ···················· | ····················· | ···················· | ···················· | ···················· |
| AA | 2416 | hCoV-19/Denmark/DCGC-18575/2020\|EPI_ISL_750055\|202 | ···················· | ···················· | ····················· | ···················· | ···················· | ···················· |
| AA | 2417 | hCoV-19/Denmark/DCGC-18578/2020\|EPI_ISL_750056\|202 | ···················· | ···················· | ····················· | ···················· | ···················· | ···················· |
| AA | 2418 | hCoV-19/Denmark/DCGC-18580/2020\|EPI_ISL_750057\|202 | ···················· | ···················· | ····················· | ···················· | ···················· | ···················· |
| AA | 2419 | hCoV-19/Denmark/DCGC-18581/2020\|EPI_ISL_750058\|202 | ···················· | ···················· | ····················· | ···················· | ···················· | ···················· |
| AA | 2420 | hCoV-19/Denmark/DCGC-18582/2020\|EPI_ISL_750059\|202 | ···················· | ···················· | ····················· | ···················· | ···················· | ···················· |
| AA | 2421 | hCoV-19/Denmark/DCGC-18583/2020\|EPI_ISL_750060\|202 | ···················· | ···················· | ····················· | ···················· | ···················· | ···················· |
| AA | 2422 | hCoV-19/Denmark/DCGC-18584/2020\|EPI_ISL_750061\|202 | ···················· | ···················· | ····················· | ···················· | ···················· | ···················· |
| AA | 2423 | hCoV-19/Denmark/DCGC-18586/2020\|EPI_ISL_750062\|202 | ···················· | ·A·················· | ····················· | ···················· | ···················· | ···················· |
| AA | 2424 | hCoV-19/Denmark/DCGC-18587/2020\|EPI_ISL_750063\|202 | ···················· | ···················· | ····················· | ···················· | ···················· | ···················· |
| AA | 2425 | hCoV-19/Denmark/DCGC-18588/2020\|EPI_ISL_750064\|202 | ···················· | ···················· | ····················· | ···················· | ···················· | ···················· |
| AA | 2426 | hCoV-19/Denmark/DCGC-18590/2020\|EPI_ISL_750065\|202 | ···················· | ···················· | ····················· | ···················· | ···················· | ···················· |
| AA | 2427 | hCoV-19/Denmark/DCGC-18591/2020\|EPI_ISL_750066\|202 | ···················· | ···················· | ····················· | ···················· | ···················· | ···················· |
| AA | 2428 | hCoV-19/Denmark/DCGC-18592/2020\|EPI_ISL_750067\|202 | ···················· | ···················· | ····················· | ···················· | ···················· | ···················· |
| AA | 2429 | hCoV-19/Denmark/DCGC-18595/2020\|EPI_ISL_750068\|202 | ···················· | ···················· | ····················· | ···················· | ···················· | ···················· |
| AA | 2430 | hCoV-19/Denmark/DCGC-18596/2020\|EPI_ISL_750069\|202 | ···················· | ···················· | ····················· | ···················· | ···················· | ···················· |
| AA | 2431 | hCoV-19/India/DL-IGIB11305069/2020\|EPI_ISL_751313\| | ···················· | ·A·················· | ····················· | ···················· | ···················· | ··A················· |
| AA | 2432 | hCoV-19/India/DL-IGIB11304690/2020\|EPI_ISL_751315\| | ···················· | ···················· | ····················· | ···················· | ···················· | ···················· |
| AA | 2433 | hCoV-19/India/DL-IGIB11305043/2020\|EPI_ISL_751316\| | ···················· | ·A·················· | ····················· | ···················· | ···················· | ··A················· |
| AA | 2434 | hCoV-19/India/DL-IGIB11304171/2020\|EPI_ISL_751317\| | ···················· | ·A·················· | ····················· | ···················· | ···················· | ··A················· |
| AA | 2435 | hCoV-19/Italy/VEN-UniVR-119/2020\|EPI_ISL_751320\|20 | ···················· | ···················· | ····················· | ···················· | ···················· | ···················· |
| AA | 2436 | hCoV-19/Italy/VEN-UniVR-130/2020\|EPI_ISL_751321\|20 | ···················· | ···················· | ···········T········· | ···················· | ···················· | ···················· |
| AA | 2437 | hCoV-19/Italy/VEN-UniVR-151/2020\|EPI_ISL_751322\|20 | ···················· | ···················· | ····················· | ···················· | ···················· | ···················· |
| AA | 2438 | hCoV-19/Italy/VEN-UniVR-29/2020\|EPI_ISL_751323\|202 | ···················· | ···················· | ····················· | ···················· | ···················· | ···················· |
| AA | 2439 | hCoV-19/Italy/VEN-UniVR-33/2020\|EPI_ISL_751324\|202 | ···················· | ···················· | ····················· | ···················· | ···················· | ···················· |
| AA | 2440 | hCoV-19/Italy/VEN-UniVR-23/2020\|EPI_ISL_751331\|202 | ···················· | ···················· | ····················· | ···················· | ···················· | ···················· |
| AA | 2441 | hCoV-19/Italy/VEN-UniVR-32/2020\|EPI_ISL_751332\|202 | ············T······· | ···················· | ····················· | ···················· | ···················· | ···················· |
| AA | 2442 | hCoV-19/Italy/VEN-UniVR-24/2020\|EPI_ISL_751333\|202 | ···················· | ···················· | ····················· | ···················· | ···················· | ···················· |
| AA | 2443 | hCoV-19/Italy/VEN-UniVR-10/2020\|EPI_ISL_751334\|202 | ···················· | ···················· | ····················· | ···················· | ···················· | ···················· |
| AA | 2444 | hCoV-19/Italy/VEN-UniVR-11/2020\|EPI_ISL_751335\|202 | ···················· | ···················· | ····················· | ···················· | ······T············· | ···················· |
| AA | 2445 | hCoV-19/Italy/VEN-UniVR-13/2020\|EPI_ISL_751336\|202 | ···················· | ···················· | ····················· | ···················· | ···················· | ···················· |
| AA | 2446 | hCoV-19/Italy/VEN-UniVR-16/2020\|EPI_ISL_751337\|202 | ···················· | ···················· | ····················· | ···················· | ···················· | ···················· |
| AA | 2447 | hCoV-19/Italy/VEN-UniVR-17/2020\|EPI_ISL_751338\|202 | ···················· | ···················· | ····················· | ···················· | ···················· | ···················· |
| AA | 2448 | hCoV-19/Italy/VEN-UniVR-18/2020\|EPI_ISL_751339\|202 | ···················· | ···················· | ····················· | ···················· | ···················· | ···················· |
| AA | 2449 | hCoV-19/Italy/VEN-UniVR-19/2020\|EPI_ISL_751340\|202 | ···················· | ···················· | ····················· | ···················· | ···················· | ···················· |
| AA | 2450 | hCoV-19/Italy/VEN-UniVR-20/2020\|EPI_ISL_751341\|202 | ···················· | ···················· | ····················· | ···················· | ···················· | ···················· |
| AA | 2451 | hCoV-19/Italy/VEN-UniVR-21/2020\|EPI_ISL_751342\|202 | ···················· | ···················· | ····················· | ···················· | ···················· | ···················· |
| AA | 2452 | hCoV-19/Italy/VEN-UniVR-22/2020\|EPI_ISL_751343\|202 | ···················· | ···················· | ····················· | ···················· | ···················· | ···················· |
| AA | 2453 | hCoV-19/Italy/VEN-UniVR-25/2020\|EPI_ISL_751344\|202 | ···················· | ···················· | ····················· | ···················· | ······T············· | ···················· |
| AA | 2454 | hCoV-19/Italy/VEN-UniVR-27/2020\|EPI_ISL_751345\|202 | ···················· | ···················· | ····················· | ···················· | ···················· | ···················· |
| AA | 2455 | hCoV-19/Italy/VEN-UniVR-31/2020\|EPI_ISL_751346\|202 | ···················· | ···················· | ····················· | ···················· | ···················· | ···················· |
| AA | 2456 | hCoV-19/Italy/VEN-UniVR-35/2020\|EPI_ISL_751347\|202 | ···················· | ···················· | ····················· | ···················· | ···················· | ···················· |
| AA | 2457 | hCoV-19/Italy/VEN-UniVR-36/2020\|EPI_ISL_751348\|202 | ···················· | ···················· | ····················· | ···················· | ···················· | ···················· |
| AA | 2458 | hCoV-19/Italy/VEN-UniVR-37/2020\|EPI_ISL_751349\|202 | ···················· | ···················· | ····················· | ···················· | ···················· | ···················· |
| AA | 2459 | hCoV-19/Italy/VEN-UniVR-41/2020\|EPI_ISL_751353\|202 | ···················· | ···················· | ····················· | ···················· | ···················· | ···················· |
| AA | 2460 | hCoV-19/Italy/VEN-UniVR-42/2020\|EPI_ISL_751354\|202 | ···················· | ···················· | ····················· | ···················· | ···················· | ···················· |
| AA | 2461 | hCoV-19/Italy/VEN-UniVR-44/2020\|EPI_ISL_751355\|202 | ···················· | ···················· | ····················· | ···················· | ···················· | ···················· |
| AA | 2462 | hCoV-19/Italy/VEN-UniVR-46/2020\|EPI_ISL_751356\|202 | ···················· | ···················· | ····················· | ···················· | ···················· | ···················· |
| AA | 2463 | hCoV-19/Italy/VEN-UniVR-58/2020\|EPI_ISL_751368\|202 | ···················· | ···················· | ····················· | ···················· | ···················· | ···················· |
| AA | 2464 | hCoV-19/Italy/VEN-UniVR-59/2020\|EPI_ISL_751369\|202 | ···················· | ···················· | ····················· | ···················· | ···················· | ···················· |
| AA | 2465 | hCoV-19/Italy/VEN-UniVR-60/2020\|EPI_ISL_751370\|202 | ···················· | ···················· | ····················· | ···················· | ···················· | ···················· |
| AA | 2466 | hCoV-19/Italy/VEN-UniVR-61/2020\|EPI_ISL_751371\|202 | ···················· | ···················· | ····················· | ···················· | ···················· | ···················· |
| AA | 2467 | hCoV-19/Italy/VEN-UniVR-62/2020\|EPI_ISL_751372\|202 | ···················· | ···················· | ····················· | ···················· | ···················· | ···················· |
| AA | 2468 | hCoV-19/Italy/VEN-UniVR-64/2020\|EPI_ISL_751373\|202 | ···················· | ···················· | ····················· | ···················· | ···················· | ···················· |
| AA | 2469 | hCoV-19/Italy/VEN-UniVR-65/2020\|EPI_ISL_751374\|202 | ···················· | ···················· | ····················· | ···················· | ···················· | ···················· |
| AA | 2470 | hCoV-19/Italy/VEN-UniVR-66/2020\|EPI_ISL_751375\|202 | ···················· | ···················· | ····················· | ···················· | ···················· | ···················· |
| AA | 2471 | hCoV-19/Italy/VEN-UniVR-67/2020\|EPI_ISL_751376\|202 | ···················· | ···················· | ····················· | ···················· | ···················· | ···················· |
| AA | 2472 | hCoV-19/Italy/VEN-UniVR-68/2020\|EPI_ISL_751377\|202 | ···················· | ···················· | ····················· | ···················· | ···················· | ···················· |
| AA | 2473 | hCoV-19/Italy/VEN-UniVR-69/2020\|EPI_ISL_751378\|202 | ···················· | ···················· | ····················· | ···················· | ···················· | ···················· |
| AA | 2474 | hCoV-19/Italy/VEN-UniVR-70/2020\|EPI_ISL_751379\|202 | ···················· | ···················· | ····················· | ···················· | ···················· | ···················· |
| AA | 2475 | hCoV-19/Italy/VEN-UniVR-71/2020\|EPI_ISL_751380\|202 | ···················· | ···················· | ····················· | ···················· | ···················· | ···················· |
| AA | 2476 | hCoV-19/Italy/VEN-UniVR-72/2020\|EPI_ISL_751381\|202 | ···················· | ···················· | ····················· | ···················· | ···················· | ···················· |
| AA | 2477 | hCoV-19/Italy/VEN-UniVR-73/2020\|EPI_ISL_751382\|202 | ···················· | ···················· | ····················· | ···················· | ···················· | ···················· |
| AA | 2478 | hCoV-19/Italy/VEN-UniVR-74/2020\|EPI_ISL_751383\|202 | ···················· | ···················· | ····················· | ···················· | ···················· | ···················· |
| AA | 2479 | hCoV-19/Italy/VEN-UniVR-75/2020\|EPI_ISL_751384\|202 | ···················· | ···················· | ····················· | ···················· | ···················· | ···················· |
| AA | 2480 | hCoV-19/Italy/VEN-UniVR-76/2020\|EPI_ISL_751385\|202 | ···················· | ···················· | ····················· | ···················· | ···················· | ···················· |
| AA | 2481 | hCoV-19/Italy/VEN-UniVR-77/2020\|EPI_ISL_751386\|202 | ···················· | ···················· | ····················· | ···················· | ···················· | ···················· |
| AA | 2482 | hCoV-19/Italy/VEN-UniVR-78/2020\|EPI_ISL_751387\|202 | ···················· | ···················· | ····················· | ···················· | ···················· | ···················· |

AA 2483 hCoV-19/Italy/VEN-UniVR-79/2020|EPI_ISL_751388|202 ···················· ···················· ···················· ···················· ···················· ····················
AA 2484 hCoV-19/Italy/VEN-UniVR-80/2020|EPI_ISL_751389|202 ···················· ···················· ···················· ···················· ···················· ····················
AA 2485 hCoV-19/Italy/VEN-UniVR-81/2020|EPI_ISL_751390|202 ···················· ···················· ···················· ···················· ···················· ····················
AA 2486 hCoV-19/Italy/VEN-UniVR-82/2020|EPI_ISL_751391|202 ···················· ···················· ···················· ···················· ···················· ····················
AA 2487 hCoV-19/Italy/VEN-UniVR-83/2020|EPI_ISL_751392|202 ···················· ···················· ···················· ···················· ···················· ····················
AA 2488 hCoV-19/Italy/VEN-UniVR-84/2020|EPI_ISL_751393|202 ···················· ···················· ···················· ···················· ···················· ····················
AA 2489 hCoV-19/Italy/VEN-UniVR-86/2020|EPI_ISL_751394|202 ···················· ···················· ···················· ···················· ···················· ····················
AA 2490 hCoV-19/Italy/VEN-UniVR-87/2020|EPI_ISL_751395|202 ···················· ···················· ···················· ···················· ···················· ····················
AA 2491 hCoV-19/Italy/VEN-UniVR-88/2020|EPI_ISL_751396|202 ···················· ···················· ···················· ···················· ···················· ····················
AA 2492 hCoV-19/Italy/VEN-UniVR-89/2020|EPI_ISL_751397|202 ···················· ···················· ···················· ···················· ···················· ····················
AA 2493 hCoV-19/Italy/VEN-UniVR-90/2020|EPI_ISL_751398|202 ···················· ···················· ···················· ···················· ···················· ····················
AA 2494 hCoV-19/Italy/VEN-UniVR-91/2020|EPI_ISL_751399|202 ···················· ···················· ···················· ···················· ···················· ····················
AA 2495 hCoV-19/USA/HI-H200076/2020|EPI_ISL_752644|2020-06 ···················· ···················· ···················· ···················· ···················· ····················
AA 2496 hCoV-19/USA/HI-H200077/2020|EPI_ISL_752645|2020-04 ···················· ···················· ···················· ···················· ···················· ····················
AA 2497 hCoV-19/USA/HI-H200078/2020|EPI_ISL_752646|2020-06 ···················· ···················· ···················· ···················· ···················· ····················
AA 2498 hCoV-19/USA/HI-H200080/2020|EPI_ISL_752647|2020-06 ···················· ···················· ···················· ···················· ···················· ····················
AA 2499 hCoV-19/USA/HI-H200081/2020|EPI_ISL_752648|2020-06 ···················· ···················· ···················· ···················· ···················· ····················
AA 2500 hCoV-19/USA/HI-H200082/2020|EPI_ISL_752649|2020-06 ···················· ···················· ···················· ···················· ···················· ····················
AA 2501 hCoV-19/USA/HI-H200083/2020|EPI_ISL_752650|2020-06 ···················· ···················· ···················· ···················· ···················· ····················
AA 2502 hCoV-19/USA/HI-H200084/2020|EPI_ISL_752651|2020-06 ···················· ···················· ···················· ···················· ···················· ····················
AA 2503 hCoV-19/USA/HI-H200085/2020|EPI_ISL_752652|2020-06 ···················· ···················· ···················· ···················· ···················· ····················
AA 2504 hCoV-19/USA/HI-H200086/2020|EPI_ISL_752653|2020-06 ···················· ···················· ···················· ···················· ···················· ····················
AA 2505 hCoV-19/USA/HI-H200087/2020|EPI_ISL_752654|2020-06 ···················· ···················· ···················· ···················· ···················· ····················
AA 2506 hCoV-19/USA/HI-H200088/2020|EPI_ISL_752655|2020-06 ···················· ···················· ···················· ···················· ···················· ····················
AA 2507 hCoV-19/USA/HI-H200089/2020|EPI_ISL_752656|2020-06 ···················· ···················· ···················· ···················· ···················· ····················
AA 2508 hCoV-19/USA/HI-H200090/2020|EPI_ISL_752657|2020-06 ···················· ···················· ···················· ···················· ···················· ····················
AA 2509 hCoV-19/USA/HI-H200091/2020|EPI_ISL_752658|2020-06 ···················· ···················· ···················· ···················· ···················· ····················
AA 2510 hCoV-19/USA/HI-H200092/2020|EPI_ISL_752659|2020-06 ···················· ···················· ···················· ···················· ···················· ····················
AA 2511 hCoV-19/USA/HI-H200093/2020|EPI_ISL_752660|2020-03 ···················· ···················· ···················· ···················· ···················· ····················
AA 2512 hCoV-19/USA/HI-H200094/2020|EPI_ISL_752661|2020-04 ···················· ···················· ···················· ···················· ···················· ····················
AA 2513 hCoV-19/USA/HI-H200097/2020|EPI_ISL_752662|2020-04 ···················· ···················· ···················· ···················· ···················· ····················
AA 2514 hCoV-19/USA/HI-H200099/2020|EPI_ISL_752663|2020-05 ···················· ···················· ···················· ···················· ···················· ····················
AA 2515 hCoV-19/USA/HI-H200100/2020|EPI_ISL_752664|2020-06 ···················· ···················· ···················· ···················· ···················· ····················
AA 2516 hCoV-19/USA/HI-H200101/2020|EPI_ISL_752665|2020-06 ···················· ···················· ···················· ···················· ···················· ····················
AA 2517 hCoV-19/USA/HI-H200102/2020|EPI_ISL_752666|2020-06 ···················· ···················· ···················· ···················· ···················· ····················
AA 2518 hCoV-19/USA/HI-H200103/2020|EPI_ISL_752667|2020-06 ···················· ···················· ···················· ···················· ···················· ····················
AA 2519 hCoV-19/USA/HI-H200105/2020|EPI_ISL_752668|2020-04 ···················· ···················· ···················· ···················· ···················· ····················
AA 2520 hCoV-19/USA/HI-H200106/2020|EPI_ISL_752669|2020-04 ···················· ···················· ···················· ···················· ···················· ····················
AA 2521 hCoV-19/USA/HI-H200107/2020|EPI_ISL_752670|2020-05 ···················· ···················· ···················· ···················· ···················· ····················
AA 2522 hCoV-19/USA/HI-H200108/2020|EPI_ISL_752671|2020-05 ···················· ···················· ···················· ···················· ···················· ····················
AA 2523 hCoV-19/USA/HI-H200109/2020|EPI_ISL_752672|2020-05 ···················· ···················· ···················· ···················· ···················· ····················
AA 2524 hCoV-19/USA/HI-H200110/2020|EPI_ISL_752673|2020-06 ···················· ···················· ···················· ···················· ···················· ····················
AA 2525 hCoV-19/USA/HI-H200111/2020|EPI_ISL_752674|2020-06 ···················· ···················· ···················· ···················· ···················· ····················
AA 2526 hCoV-19/USA/HI-H200112/2020|EPI_ISL_752675|2020-06 ···················· ···················· ···················· ···················· ···················· ····················
AA 2527 hCoV-19/USA/HI-H200113/2020|EPI_ISL_752676|2020-06 ···················· ···················· ···················· ···················· ···················· ····················
AA 2528 hCoV-19/USA/HI-H200114/2020|EPI_ISL_752677|2020-06 ···················· ···················· ···················· ···················· ···················· ····················
AA 2529 hCoV-19/USA/HI-H200115/2020|EPI_ISL_752678|2020-06 ···················· ···················· ···················· ···················· ···················· ····················
AA 2530 hCoV-19/USA/HI-H200116/2020|EPI_ISL_752679|2020-06 ···················· ···················· ···················· ···················· ···················· ····················
AA 2531 hCoV-19/USA/HI-H200121/2020|EPI_ISL_752684|2020-06 ···················· ···················· ···················· ···················· ···················· ····················
AA 2532 hCoV-19/USA/HI-H200122/2020|EPI_ISL_752685|2020-06 ···················· ···················· ···················· ···················· ···················· ····················
AA 2533 hCoV-19/USA/HI-H200123/2020|EPI_ISL_752686|2020-06 ···················· ···················· ···················· ···················· ···················· ····················
AA 2534 hCoV-19/USA/HI-H200124/2020|EPI_ISL_752687|2020-06 ···················· ···················· ···················· ···················· ···················· ····················
AA 2535 hCoV-19/USA/HI-H200136/2020|EPI_ISL_752699|2020-07 ···················· ···················· ···················· ···················· ···················· ····················
AA 2536 hCoV-19/Germany/DE-BE-ChVir-W1638-6728/2020|EPI_IS ···················· ·A·················· ···················· ···················· ···················· ····················
AA 2537 hCoV-19/Germany/DE-BE-ChVir-W1639-6740/2020|EPI_IS ···················· ···················· ···················· ···················· ··A················· ····················
AA 2538 hCoV-19/Germany/DE-BE-ChVir-W1642-6757/2020|EPI_IS ···················· ···················· ···················· ···················· ···················· ····················
AA 2539 hCoV-19/Germany/DE-BE-ChVir-W1644-6714/2020|EPI_IS ···················· ···················· ···················· ···················· ···················· ····················
AA 2540 hCoV-19/Germany/DE-BE-ChVir-W1645-6781/2020|EPI_IS ···················· ·A·················· ···················· ···················· ···················· ····················
AA 2541 hCoV-19/Germany/DE-BE-ChVir-W1647-6745/2020|EPI_IS ···················· ···················A ···················· ···················· ···················· ···················· 1
AA 2542 hCoV-19/Germany/DE-BE-ChVir-W1649-7416/2020|EPI_IS ···················· ···················· ···················· ···················· ···················· ····················
AA 2543 hCoV-19/Germany/DE-BE-ChVir-W1651-8237/2020|EPI_IS ···················· ···················· ···················· ···················· ···················· ····················
AA 2544 hCoV-19/Germany/DE-BE-ChVir-W1652-8241/2020|EPI_IS ···················· ···················· ···················· ···················· ···················· ····················
AA 2545 hCoV-19/Germany/DE-BE-ChVir-W1688-4369/2020|EPI_IS ···················· ···················· ···················· ···················· ···················· ····················
AA 2546 hCoV-19/Germany/DE-BE-ChVir-W1690-8217/2020|EPI_IS ···················· ···················· ···················· ···················· ···················· ····················
AA 2547 hCoV-19/Germany/DE-BE-ChVir-W1691-8296/2020|EPI_IS ···················· ···················· ···················· ···················· ···················· ····················
AA 2548 hCoV-19/Germany/DE-BE-ChVir-W1692-8235/2020|EPI_IS ···················· ···················· ···················· ···················· ···················· ····················
AA 2549 hCoV-19/Germany/DE-BE-ChVir-W1693-4382/2020|EPI_IS ···················· ···················· ···················· ···················· ···················· ····················
AA 2550 hCoV-19/Germany/DE-BE-ChVir-W1694-8256/2020|EPI_IS ···················· ···················· ···················· ···················· ···················· ····················
AA 2551 hCoV-19/Germany/DE-BE-ChVir-W1695-8202/2020|EPI_IS ···················· ···················· ···················· ···················· ···················· ····················
AA 2552 hCoV-19/Germany/DE-BE-ChVir-W1696-8291/2020|EPI_IS ···················· ···················· ···················· ···················· ···················· ····················
AA 2553 hCoV-19/Germany/DE-BE-ChVir-W1697-8293/2020|EPI_IS ···················· ···················· ···················· ···················· ···················· ····················
AA 2554 hCoV-19/Germany/DE-BE-ChVir-W1698-8200/2020|EPI_IS ···················· ···················· ···················· ···················· ···················· ····················
AA 2555 hCoV-19/Germany/DE-BE-ChVir-W1700-6762/2020|EPI_IS ···················· ···················· ···················· ···················· ···················· ····················
AA 2556 hCoV-19/Germany/DE-BE-ChVir-W1703-6771/2020|EPI_IS ···················· ···················· ···················· ···················· ···················· ····················
AA 2557 hCoV-19/Germany/DE-BE-ChVir-W1704-4391/2020|EPI_IS ···················· ···················· ···················· ···················· ···················· ····················
AA 2558 hCoV-19/Germany/DE-BE-ChVir-W1706-8239/2020|EPI_IS ···················· ···················· ···················· ···················· ···················· ····················
AA 2559 hCoV-19/Germany/DE-BE-ChVir-W1708-6787/2020|EPI_IS ···················· ···················· ···················· ···················· ···················· ····················
AA 2560 hCoV-19/Germany/DE-BE-ChVir-W1709-8206/2020|EPI_IS ···················· ···················· ···················· ···················· ···················· ····················
AA 2561 hCoV-19/Denmark/DCGC-19645/2020|EPI_ISL_749080|202 ···················· ···················· ···················· ···················· ···················· ····················
AA 2562 hCoV-19/Denmark/DCGC-19647/2020|EPI_ISL_749081|202 ···················· ···················· ···················· ···················· ···················· ····················

| | | | | | | | |
|---|---|---|---|---|---|---|---|
| AA | 2563 hCoV-19/Denmark/DCGC-19648/2020\|EPI_ISL_749082\|202 | ···················· | ···················· | ···················· | ···················· | ···················· | ···················· |
| AA | 2564 hCoV-19/Denmark/DCGC-19649/2020\|EPI_ISL_749083\|202 | ···················· | ···················· | ···················· | ···················· | ···················· | ···················· |
| AA | 2565 hCoV-19/Denmark/DCGC-19651/2020\|EPI_ISL_749084\|202 | ···················· | ·A·················· | ···················· | ···················· | ···················· | ··A················· |
| AA | 2566 hCoV-19/Denmark/DCGC-19652/2020\|EPI_ISL_749085\|202 | ···················· | ···················· | ···················· | ···················· | ···················· | ···················· |
| AA | 2567 hCoV-19/Denmark/DCGC-19653/2020\|EPI_ISL_749086\|202 | ···················· | ···················· | ···················· | ···················· | ···················· | ···················· |
| AA | 2568 hCoV-19/Denmark/DCGC-19655/2020\|EPI_ISL_749087\|202 | ···················· | ···················· | ···················· | ···················· | ···················· | ···················· |
| AA | 2569 hCoV-19/Denmark/DCGC-19656/2020\|EPI_ISL_749088\|202 | ···················· | ···················· | ···················· | ···················· | ···················· | ···················· |
| AA | 2570 hCoV-19/Denmark/DCGC-19657/2020\|EPI_ISL_749089\|202 | ···················· | ···················· | ···················· | ···················· | ···················· | ···················· |
| AA | 2571 hCoV-19/Denmark/DCGC-19658/2020\|EPI_ISL_749090\|202 | ···················· | ···················· | ···················· | ···················· | ···················· | ···················· |
| AA | 2572 hCoV-19/Denmark/DCGC-19659/2020\|EPI_ISL_749091\|202 | ···················· | ···················· | ···················· | ···················· | ···················· | ···················· |
| AA | 2573 hCoV-19/Denmark/DCGC-19660/2020\|EPI_ISL_749092\|202 | ···················· | ···················· | ···················· | ···················· | ···················· | ···················· |
| AA | 2574 hCoV-19/Denmark/DCGC-19661/2020\|EPI_ISL_749093\|202 | ···················· | ···················· | ···················· | ···················· | ···················· | ···················· |
| AA | 2575 hCoV-19/Denmark/DCGC-19662/2020\|EPI_ISL_749094\|202 | ···················· | ···················· | ···················· | ···················· | ···················· | ···················· |
| AA | 2576 hCoV-19/Denmark/DCGC-19664/2020\|EPI_ISL_749095\|202 | ···················· | ·A·················· | ···················· | ···················· | ···················· | ··A················· |
| AA | 2577 hCoV-19/Denmark/DCGC-19665/2020\|EPI_ISL_749096\|202 | ···················· | ···················· | ···················· | ···················· | ···················· | ···················· |
| AA | 2578 hCoV-19/Denmark/DCGC-19667/2020\|EPI_ISL_749097\|202 | ···················· | ···················· | ···················· | ···················· | ···················· | ···················· |
| AA | 2579 hCoV-19/Denmark/DCGC-19668/2020\|EPI_ISL_749098\|202 | ···················· | ···················· | ···················· | ···················· | ···················· | ···················· |
| AA | 2580 hCoV-19/Denmark/DCGC-19669/2020\|EPI_ISL_749099\|202 | ···················· | ···················· | ···················· | ···················· | ···················· | ···················· |
| AA | 2581 hCoV-19/Denmark/DCGC-18525/2020\|EPI_ISL_750020\|202 | ···················· | ···················· | ···················· | ···················· | ···················· | ···················· |
| AA | 2582 hCoV-19/Denmark/DCGC-18526/2020\|EPI_ISL_750021\|202 | ···················· | ·A·················· | ···················· | ···················· | ···················· | ··A················· |
| AA | 2583 hCoV-19/Denmark/DCGC-18531/2020\|EPI_ISL_750023\|202 | ···················· | ···················· | ···················· | ···················· | ···················· | ···················· |
| AA | 2584 hCoV-19/Denmark/DCGC-18597/2020\|EPI_ISL_750070\|202 | ···················· | ···················· | ···················· | ···················· | ···················· | ···················· |
| AA | 2585 hCoV-19/Denmark/DCGC-18598/2020\|EPI_ISL_750071\|202 | ···················· | ·A·················· | ···················· | ···················· | ···················· | ··A················· |
| AA | 2586 hCoV-19/Denmark/DCGC-18599/2020\|EPI_ISL_750072\|202 | ···················· | ···················· | ···················· | ···················· | ···················· | ···················· |
| AA | 2587 hCoV-19/Denmark/DCGC-18601/2020\|EPI_ISL_750073\|202 | ···················· | ···················· | ···················· | ···················· | ···················· | ···················· |
| AA | 2588 hCoV-19/Denmark/DCGC-18602/2020\|EPI_ISL_750074\|202 | ···················· | ···················· | ···················· | ···················· | ···················· | ···················· |
| AA | 2589 hCoV-19/Denmark/DCGC-18606/2020\|EPI_ISL_750075\|202 | ···················· | ···················· | ···················· | ···················· | ···················· | ···················· |
| AA | 2590 hCoV-19/Denmark/DCGC-18608/2020\|EPI_ISL_750076\|202 | ···················· | ···················· | ···················· | ···················· | ···················· | ···················· |
| AA | 2591 hCoV-19/Denmark/DCGC-18610/2020\|EPI_ISL_750077\|202 | ···················· | ···················· | ···················· | ···················· | ···················· | ···················· |
| AA | 2592 hCoV-19/Denmark/DCGC-18611/2020\|EPI_ISL_750078\|202 | ···················· | ···················· | ···················· | ···················· | ···················· | ···················· |
| AA | 2593 hCoV-19/Denmark/DCGC-18612/2020\|EPI_ISL_750079\|202 | ···················· | ···················· | ···················· | ···················· | ···················· | ···················· |
| AA | 2594 hCoV-19/Denmark/DCGC-18613/2020\|EPI_ISL_750080\|202 | ···················· | ···················· | ···················· | ···················· | ···················· | ···················· |
| AA | 2595 hCoV-19/Denmark/DCGC-18615/2020\|EPI_ISL_750081\|202 | ···················· | ···················· | ···················· | ···················· | ···················· | ···················· |
| AA | 2596 hCoV-19/Denmark/DCGC-18616/2020\|EPI_ISL_750082\|202 | ···················· | ···················· | ···················· | ···················· | ···················· | ···················· |
| AA | 2597 hCoV-19/Denmark/DCGC-18617/2020\|EPI_ISL_750083\|202 | ···················· | ······A············· | ···················· | ···················· | ···················· | ···················· |
| AA | 2598 hCoV-19/Denmark/DCGC-18618/2020\|EPI_ISL_750084\|202 | ···················· | ···················· | ···················· | ···················· | ···················· | ···················· |
| AA | 2599 hCoV-19/Denmark/DCGC-18621/2020\|EPI_ISL_750085\|202 | ···················· | ···················· | ···················· | ···················· | ···················· | ···················· |
| AA | 2600 hCoV-19/Denmark/DCGC-18622/2020\|EPI_ISL_750086\|202 | ···················· | ···················· | ···················· | ···················· | ···················· | ···················· |
| AA | 2601 hCoV-19/Denmark/DCGC-18623/2020\|EPI_ISL_750087\|202 | ···················· | ···················· | ···················· | ···················· | ···················· | ···················· |
| AA | 2602 hCoV-19/Denmark/DCGC-18624/2020\|EPI_ISL_750088\|202 | ···················· | ···················· | ···················· | ···················· | ···················· | ···················· |
| AA | 2603 hCoV-19/Denmark/DCGC-18625/2020\|EPI_ISL_750089\|202 | ···················· | ···················· | ···················· | ···················· | ···················· | ···················· |
| AA | 2604 hCoV-19/Denmark/DCGC-18628/2020\|EPI_ISL_750090\|202 | ···················· | ···················· | ···················· | ···················· | ···················· | ···················· |
| AA | 2605 hCoV-19/Denmark/DCGC-18629/2020\|EPI_ISL_750091\|202 | ···················· | ···················· | ···················· | ···················· | ···················· | ···················· |
| AA | 2606 hCoV-19/Denmark/DCGC-18630/2020\|EPI_ISL_750092\|202 | ···················· | ···················· | ···················· | ···················· | ···················· | ···················· |
| AA | 2607 hCoV-19/Denmark/DCGC-18634/2020\|EPI_ISL_750093\|202 | ···················· | ···················· | ···················· | ···················· | ···················· | ···················· |
| AA | 2608 hCoV-19/Denmark/DCGC-18637/2020\|EPI_ISL_750094\|202 | ···················· | ···················· | ···················· | ···················· | ···················· | ···················· |
| AA | 2609 hCoV-19/Denmark/DCGC-18638/2020\|EPI_ISL_750095\|202 | ···················· | ···················· | ···················· | ···················· | ···················· | ···················· |
| AA | 2610 hCoV-19/Denmark/DCGC-18642/2020\|EPI_ISL_750096\|202 | ···················· | ············G······· | ···················· | ···················· | ···················· | ···················· |
| AA | 2611 hCoV-19/Denmark/DCGC-18643/2020\|EPI_ISL_750097\|202 | ···················· | ···················· | ···················· | ···················· | ···················· | ···················· |
| AA | 2612 hCoV-19/Denmark/DCGC-18644/2020\|EPI_ISL_750098\|202 | ···················· | ···················· | ···················· | ···················· | ···················· | ···················· |
| AA | 2613 hCoV-19/Denmark/DCGC-18646/2020\|EPI_ISL_750099\|202 | ···················· | ···················· | ···················· | ···················· | ···················· | ···················· |
| AA | 2614 hCoV-19/Italy/VEN-UniVR-38/2020\|EPI_ISL_751350\|202 | ···················· | ···················· | ···················· | ···················· | ···················· | ···················· |
| AA | 2615 hCoV-19/Italy/VEN-UniVR-39/2020\|EPI_ISL_751351\|202 | ···················· | ···················· | ···················· | ···················· | ···················· | ···················· |
| AA | 2616 hCoV-19/Italy/VEN-UniVR-40/2020\|EPI_ISL_751352\|202 | ···················· | ·A·················· | ···················· | ···················· | ···················· | ···················· |
| AA | 2617 hCoV-19/Italy/VEN-UniVR-92/2020\|EPI_ISL_751400\|202 | ···················· | ···················· | ···················· | ···················· | ···················· | ···················· |
| AA | 2618 hCoV-19/Italy/VEN-UniVR-93/2020\|EPI_ISL_751401\|202 | ···················· | ···················· | ···················· | ···················· | ···················· | ···················· |
| AA | 2619 hCoV-19/Italy/VEN-UniVR-94/2020\|EPI_ISL_751402\|202 | ···················· | ···················· | ···················· | ···················· | ···················· | ···················· |
| AA | 2620 hCoV-19/Italy/VEN-UniVR-95/2020\|EPI_ISL_751403\|202 | ···················· | ···················· | ···················· | ···················· | ···················· | ···················· |
| AA | 2621 hCoV-19/Italy/VEN-UniVR-98/2020\|EPI_ISL_751404\|202 | ···················· | ···················· | ···················· | ···················· | ···················· | ···················· |
| AA | 2622 hCoV-19/Italy/VEN-UniVR-99/2020\|EPI_ISL_751405\|202 | ···················· | ···················· | ···················· | ···················· | ···················· | ···················· |
| AA | 2623 hCoV-19/Italy/VEN-UniVR-100/2020\|EPI_ISL_751406\|20 | ···················· | ···················· | ···················· | ···················· | ···················· | ···················· |
| AA | 2624 hCoV-19/Italy/VEN-UniVR-101/2020\|EPI_ISL_751407\|20 | ···················· | ···················· | ···················· | ···················· | ···················· | ···················· |
| AA | 2625 hCoV-19/Italy/VEN-UniVR-103/2020\|EPI_ISL_751408\|20 | ···················· | ···················· | ···················· | ···················· | ···················· | ···················· |
| AA | 2626 hCoV-19/Italy/VEN-UniVR-104/2020\|EPI_ISL_751409\|20 | ···················· | ···················· | ···················· | ···················· | ···················· | ···················· |
| AA | 2627 hCoV-19/Italy/VEN-UniVR-105/2020\|EPI_ISL_751410\|20 | ···················· | ···················· | ···················· | ···················· | ···················· | ···················· |
| AA | 2628 hCoV-19/Italy/VEN-UniVR-106/2020\|EPI_ISL_751411\|20 | ···················· | ···················· | ···················· | ···················· | ···················· | ···················· |
| AA | 2629 hCoV-19/Italy/VEN-UniVR-117/2020\|EPI_ISL_751418\|20 | ···················· | ···················· | ···················· | ···················· | ···················· | ···················· |
| AA | 2630 hCoV-19/Italy/VEN-UniVR-118/2020\|EPI_ISL_751419\|20 | ···················· | ···················· | ···················· | ···················· | ···················· | ···················· |
| AA | 2631 hCoV-19/USA/HI-H200117/2020\|EPI_ISL_752680\|2020-06 | ···················· | ···················· | ···················· | ···················· | ···················· | ···················· |
| AA | 2632 hCoV-19/USA/HI-H200118/2020\|EPI_ISL_752681\|2020-06 | ···················· | ···················· | ···················· | ···················· | ···················· | ···················· |
| AA | 2633 hCoV-19/USA/HI-H200119/2020\|EPI_ISL_752682\|2020-06 | ···················· | ···················· | ···················· | ···················· | ···················· | ···················· |
| AA | 2634 hCoV-19/USA/HI-H200120/2020\|EPI_ISL_752683\|2020-06 | ···················· | ···················· | ···················· | ···················· | ···················· | ···················· |
| AA | 2635 hCoV-19/USA/HI-H200159/2020\|EPI_ISL_752721\|2020-07 | ···················· | ···················· | ···················· | ···················· | ···················· | ···················· |
| AA | 2636 hCoV-19/USA/HI-H200160/2020\|EPI_ISL_752722\|2020-07 | ···················· | ···················· | ···················· | ···················· | ···················· | ···················· |
| AA | 2637 hCoV-19/USA/HI-H200162/2020\|EPI_ISL_752723\|2020-07 | ···················· | ·A·················· | ···················· | ···················· | ···················· | ···················· |
| AA | 2638 hCoV-19/USA/HI-H200163/2020\|EPI_ISL_752724\|2020-07 | ···················· | ·A·················· | ···················· | ···················· | ···················· | ···················· |
| AA | 2639 hCoV-19/USA/HI-H200164/2020\|EPI_ISL_752725\|2020-07 | ···················· | ·A·················· | ···················· | ···················· | ···················· | ···················· |
| AA | 2640 hCoV-19/USA/HI-H200165/2020\|EPI_ISL_752726\|2020-07 | ···················· | ···················· | ···················· | ···················· | ···················· | ···················· |
| AA | 2641 hCoV-19/USA/HI-H200166/2020\|EPI_ISL_752727\|2020-07 | ···················· | ···················· | ···················· | ···················· | ···················· | ···················· |
| AA | 2642 hCoV-19/USA/HI-H200167/2020\|EPI_ISL_752728\|2020-07 | ···················· | ···················· | ···················· | ···················· | ···················· | ···················· |

| | | | | | | | |
|---|---|---|---|---|---|---|---|
| AA | 2643 hCoV-19/USA/HI-H200168/2020\|EPI_ISL_752729\|2020-07 | ···················· | ···················· | ···················· | ···················· | ···················· | ···················· |
| AA | 2644 hCoV-19/USA/HI-H200169/2020\|EPI_ISL_752730\|2020-07 | ···················· | ···················· | ···················· | ···················· | ···················· | ···················· |
| AA | 2645 hCoV-19/USA/HI-H200170/2020\|EPI_ISL_752731\|2020-07 | ···················· | ·A·················· | ···················· | ···················· | ···················· | ···················· |
| AA | 2646 hCoV-19/USA/HI-H200171/2020\|EPI_ISL_752732\|2020-07 | ···················· | ·A·················· | ···················· | ···················· | ···················· | ···················· |
| AA | 2647 hCoV-19/USA/HI-H200173/2020\|EPI_ISL_752733\|2020-07 | ···················· | ·A·················· | ···················· | ···················· | ···················· | ···················· |
| AA | 2648 hCoV-19/USA/HI-H200174/2020\|EPI_ISL_752734\|2020-07 | ···················· | ·A·················· | ···················· | ···················· | ···················· | ···················· |
| AA | 2649 hCoV-19/USA/HI-H200175/2020\|EPI_ISL_752735\|2020-07 | ···················· | ·A·················· | ···················· | ···················· | ···················· | ···················· |
| AA | 2650 hCoV-19/USA/HI-H200176/2020\|EPI_ISL_752736\|2020-07 | ···················· | ·A·················· | ···················· | ···················· | ···················· | ···················· |
| AA | 2651 hCoV-19/USA/HI-H200177/2020\|EPI_ISL_752737\|2020-07 | ···················· | ·A·················· | ···················· | ···················· | ···················· | ···················· |
| AA | 2652 hCoV-19/USA/HI-H200178/2020\|EPI_ISL_752738\|2020-07 | ···················· | ·A·················· | ···················· | ···················· | ···················· | ···················· |
| AA | 2653 hCoV-19/USA/HI-H200179/2020\|EPI_ISL_752739\|2020-07 | ···················· | ·A·················· | ···················· | ···················· | ···················· | ···················· |
| AA | 2654 hCoV-19/USA/HI-H200180/2020\|EPI_ISL_752740\|2020-07 | ···················· | ·A·················· | ···················· | ···················· | ···················· | ···················· |
| AA | 2655 hCoV-19/USA/HI-H200181/2020\|EPI_ISL_752741\|2020-07 | ···················· | ·A·················· | ···················· | ···················· | ···················· | ···················· |
| AA | 2656 hCoV-19/USA/HI-H200182/2020\|EPI_ISL_752742\|2020-07 | ···················· | ·A·················· | ···················· | ···················· | ···················· | ···················· |
| AA | 2657 hCoV-19/USA/HI-H200190/2020\|EPI_ISL_752749\|2020-08 | ···················· | ·A·················· | ···················· | ···················· | ···················· | ···················· |
| AA | 2658 hCoV-19/USA/WA-UW-46899/2020\|EPI_ISL_756200\|2020-1 | ···················· | ···················· | ···················· | ···················· | ···················· | ···················· |
| AA | 2659 hCoV-19/USA/WA-UW-46903/2020\|EPI_ISL_756201\|2020-1 | ···················· | ···················· | ···················· | ···················· | ···················· | ···················· |
| AA | 2660 hCoV-19/USA/WA-UW-46905/2020\|EPI_ISL_756202\|2020-1 | ···················· | ···················· | ···················· | ···················· | ···················· | ···················· |
| AA | 2661 hCoV-19/USA/WA-UW-46906/2020\|EPI_ISL_756203\|2020-1 | ···················· | ·A·················· | ···················· | ············A······· | ···················· | ···················· |
| AA | 2662 hCoV-19/USA/WA-UW-46923/2020\|EPI_ISL_756205\|2020-1 | ···················· | ···················· | ···················· | ···················· | ···················· | ···················· |
| AA | 2663 hCoV-19/USA/WA-UW-46934/2020\|EPI_ISL_756206\|2020-1 | ···················· | ···················· | ···················· | ···················· | ···················· | ···················· |
| AA | 2664 hCoV-19/USA/WA-UW-46937/2020\|EPI_ISL_756207\|2020-1 | ···················· | ···················· | ···················· | ···················· | ···················· | ···················· |
| AA | 2665 hCoV-19/USA/WA-UW-46945/2020\|EPI_ISL_756209\|2020-1 | ···················· | ···················· | ···················· | ···················· | ···················· | ···················· |
| AA | 2666 hCoV-19/USA/WA-UW-46947/2020\|EPI_ISL_756210\|2020-1 | ···················· | ···················· | ···················· | ···················· | ···················· | ···················· |
| AA | 2667 hCoV-19/USA/WA-UW-46961/2020\|EPI_ISL_756212\|2020-1 | ···················· | ···················· | ···················· | ···················· | ···················· | ···················· |
| AA | 2668 hCoV-19/USA/WA-UW-46966/2020\|EPI_ISL_756213\|2020-1 | ···················· | ···················· | ···················· | ···················· | ···················· | ···················· |
| AA | 2669 hCoV-19/USA/WA-UW-46976/2020\|EPI_ISL_756214\|2020-1 | ···················· | ···················· | ···················· | ···················· | ···················· | ···················· |
| AA | 2670 hCoV-19/USA/WA-UW-46997/2020\|EPI_ISL_756215\|2020-1 | ···················· | ·A·················· | ···················· | ···················· | ···················· | ···················· |
| AA | 2671 hCoV-19/USA/WA-UW-46998/2020\|EPI_ISL_756216\|2020-1 | ···················· | ···················· | ···················· | ···················· | ···················· | ···················· |
| AA | 2673 hCoV-19/USA/WA-UW-47030/2020\|EPI_ISL_756218\|2020-1 | ···················· | ···················· | ···················· | ···················· | ···················· | ···················· |
| AA | 2674 hCoV-19/USA/WA-UW-47031/2020\|EPI_ISL_756219\|2020-1 | ···················· | ···················· | ···················· | ···················· | ···················· | ···················· |
| AA | 2675 hCoV-19/USA/WA-UW-47034/2020\|EPI_ISL_756220\|2020-1 | ···················· | ···················· | ···················· | ···················· | ···················· | ···················· |
| AA | 2676 hCoV-19/USA/WA-UW-47049/2020\|EPI_ISL_756221\|2020-1 | ···················· | ···················· | ···················· | ···················· | ···················· | ···················· |
| AA | 2677 hCoV-19/USA/WA-UW-47079/2020\|EPI_ISL_756223\|2020-1 | ···················· | ·A·················· | ···················· | ···················· | ···················· | ···················· |
| AA | 2678 hCoV-19/USA/WA-UW-47081/2020\|EPI_ISL_756224\|2020-1 | ···················· | ···················· | ···················· | ···················· | ···················· | ···················· |
| AA | 2679 hCoV-19/USA/WA-UW-47095/2020\|EPI_ISL_756226\|2020-1 | ···················· | ·A·················· | ···················· | ···················· | ···················· | ···················· |
| AA | 2680 hCoV-19/USA/WA-UW-47100/2020\|EPI_ISL_756227\|2020-1 | ···················· | ·A·················· | ···················· | ···················· | ···················· | ···················· |
| AA | 2681 hCoV-19/USA/WA-UW-47120/2020\|EPI_ISL_756228\|2020-1 | ···················· | ·A·················· | ···················· | ···················· | ···················· | ···················· |
| AA | 2682 hCoV-19/USA/WA-UW-47123/2020\|EPI_ISL_756229\|2020-1 | ···················· | ···················· | ···················· | ···················· | ···················· | ···················· |
| AA | 2683 hCoV-19/USA/WA-UW-47127/2020\|EPI_ISL_756230\|2020-1 | ···················· | ···················· | ···················· | ···················· | ···················· | ···················· |
| AA | 2684 hCoV-19/USA/WA-UW-47130/2020\|EPI_ISL_756231\|2020-1 | ···················· | ·A·················· | ···················· | ···················· | ···················· | ···················· |
| AA | 2685 hCoV-19/USA/WA-UW-47134/2020\|EPI_ISL_756233\|2020-1 | ···················· | ···················· | ···················· | ···················· | ···················· | ···················· |
| AA | 2686 hCoV-19/USA/WA-UW-47136/2020\|EPI_ISL_756234\|2020-1 | ···················· | ···················· | ···················· | ···················· | ···················· | ···················· |
| AA | 2687 hCoV-19/USA/WA-UW-47139/2020\|EPI_ISL_756235\|2020-1 | ···················· | ···················· | ···················· | ···················· | ···················· | ···················· |
| AA | 2688 hCoV-19/USA/WA-UW-47141/2020\|EPI_ISL_756236\|2020-1 | ···················· | ·A·················· | ···················· | ···················· | ···················· | ···················· |
| AA | 2689 hCoV-19/USA/WA-UW-47144/2020\|EPI_ISL_756237\|2020-1 | ···················· | ···················· | ···················· | ···················· | ···················· | ···················· |
| AA | 2690 hCoV-19/USA/WA-UW-47147/2020\|EPI_ISL_756238\|2020-1 | ···················· | ···················· | ···················· | ···················· | ···················· | ···················· |
| AA | 2691 hCoV-19/USA/WA-UW-47148/2020\|EPI_ISL_756239\|2020-1 | ···················· | ···················· | ···················· | ···················· | ···················· | ···················· |
| AA | 2692 hCoV-19/USA/WA-UW-47151/2020\|EPI_ISL_756240\|2020-1 | ···················· | ···················· | ···················· | ···················· | ···················· | ···················· |
| AA | 2693 hCoV-19/USA/WY-WPHL-20157765/2020\|EPI_ISL_756241\|2 | ···················· | ···················· | ···················· | ···················· | ···················· | ···················· |
| AA | 2694 hCoV-19/USA/WY-WPHL-20157841/2020\|EPI_ISL_756242\|2 | ···················· | ···················· | ···················· | ···················· | ···················· | ···················· |
| AA | 2695 hCoV-19/USA/WY-WPHL-20158237/2020\|EPI_ISL_756243\|2 | ···················· | ···················· | ···················· | ···················· | ···················· | ···················· |
| AA | 2696 hCoV-19/USA/WY-WPHL-20158467/2020\|EPI_ISL_756245\|2 | ···················· | ···················· | ···················· | ···················· | ···················· | ···················· |
| AA | 2697 hCoV-19/USA/WY-WPHL-20158480/2020\|EPI_ISL_756246\|2 | ···················· | ···················· | ···················· | ···················· | ···················· | ···················· |
| AA | 2698 hCoV-19/Finland/12KS4D4/2020\|EPI_ISL_756247\|2020-0 | ···················· | ···················· | ···················· | ···················· | ···················· | ···················· |
| AA | 2699 hCoV-19/Finland/17ES5B7/2020\|EPI_ISL_756248\|2020-0 | ···················· | ···················· | ···················· | ···················· | ···················· | ···················· |
| AA | 2700 hCoV-19/Finland/17SS10A7/2020\|EPI_ISL_756249\|2020- | ···················· | ···················· | ···················· | ···················· | ···················· | ···················· |
| AA | 2701 hCoV-19/Finland/17SS13C11/2020\|EPI_ISL_756250\|2020 | ···················· | ·A·················· | ···················· | ···················· | ···················· | ··A················· |
| AA | 2702 hCoV-19/Finland/17SS20F1/2020\|EPI_ISL_756251\|2020- | ···················· | ···················· | ···················· | ···················· | ···················· | ···················· |
| AA | 2703 hCoV-19/Finland/17SS4A7/2020\|EPI_ISL_756252\|2020-0 | ···················· | ···················· | ···················· | ···················· | ···················· | ···················· |
| AA | 2704 hCoV-19/USA/WY-WPHL-20158485/2020\|EPI_ISL_756253\|2 | ···················· | ···················· | ···················· | ···················· | ···················· | ···················· |
| AA | 2705 hCoV-19/USA/WY-WPHL-20158489/2020\|EPI_ISL_756254\|2 | ···················· | ···················· | ···················· | ···················· | ···················· | ···················· |
| AA | 2706 hCoV-19/USA/WY-WPHL-20158626/2020\|EPI_ISL_756256\|2 | ···················· | ···················· | ···················· | ···················· | ···················· | ···················· |
| AA | 2707 hCoV-19/USA/WY-WPHL-20158682/2020\|EPI_ISL_756258\|2 | ···················· | ···················· | ···················· | ···················· | ···················· | ···················· |
| AA | 2708 hCoV-19/USA/WY-WPHL-20169758/2020\|EPI_ISL_756260\|2 | ···················· | ···················· | ···················· | ···················· | ···················· | ···················· |
| AA | 2709 hCoV-19/USA/WY-WPHL-20170351/2020\|EPI_ISL_756268\|2 | ···················· | ···················· | ···················· | ···················· | ···················· | ···················· |
| AA | 2710 hCoV-19/USA/WY-WPHL-20170352/2020\|EPI_ISL_756269\|2 | ···················· | ···················· | ···················· | ···················· | ···················· | ···················· |
| AA | 2711 hCoV-19/Finland/18ES16AE7/2020\|EPI_ISL_756282\|2020 | ···················· | ···················· | ···················· | ···················· | ···················· | ···················· |
| AA | 2712 hCoV-19/Finland/18ES5AH5/2020\|EPI_ISL_756283\|2020- | ···················· | ···················· | ···················· | ···················· | ···················· | ···················· |
| AA | 2713 hCoV-19/Finland/18ES6AH10/2020\|EPI_ISL_756284\|2020 | ···················· | ·A·················· | ···················· | ···················· | ···················· | ··A················· |
| AA | 2714 hCoV-19/Finland/18LS10AC5/2020\|EPI_ISL_756285\|2020 | ···················· | ·A·················· | ···················· | ···················· | ···················· | ··A················· |
| AA | 2715 hCoV-19/Finland/18LS2AB3/2020\|EPI_ISL_756286\|2020- | ···················· | ···················· | ···················· | ···················· | ···················· | ···················· |
| AA | 2716 hCoV-19/Finland/18LS2AB7/2020\|EPI_ISL_756287\|2020- | ···················· | ·A·················· | ···················· | ···················· | ···················· | ··A················· |
| AA | 2717 hCoV-19/Finland/18SS11E3/2020\|EPI_ISL_756288\|2020- | ···················· | ···················· | ···················· | ···················· | ···················· | ···················· |
| AA | 2718 hCoV-19/Finland/18SS12E4/2020\|EPI_ISL_756289\|2020- | ···················· | ·A·················· | ···················· | ···················· | ···················· | ··A················· |
| AA | 2719 hCoV-19/Denmark/DCGC-22359/2020\|EPI_ISL_757510\|202 | ···················· | ···················· | ···················· | ···················· | ···················· | ···················· |
| AA | 2720 hCoV-19/Denmark/DCGC-22360/2020\|EPI_ISL_757511\|202 | ···················· | ···················· | ···················· | ···················· | ···················· | ···················· |
| AA | 2721 hCoV-19/Denmark/DCGC-22361/2020\|EPI_ISL_757512\|202 | ···················· | ···················· | ···················· | ···················· | ···················· | ···················· |
| AA | 2722 hCoV-19/Denmark/DCGC-22362/2020\|EPI_ISL_757513\|202 | ···················· | ···················· | ···················· | ···················· | ···················· | ···················· |
| AA | 2723 hCoV-19/Denmark/DCGC-22363/2020\|EPI_ISL_757514\|202 | ···················· | ···················· | ···················· | ···················· | ···················· | ···················· |

| | | | | | | | |
|---|---|---|---|---|---|---|---|
| AA | 2724 hCoV-19/Denmark/DCGC-22364/2020\|EPI_ISL_757515\|202 | ···················· | ···················· | ····················· | ···················· | ···················· | ···················· |
| AA | 2725 hCoV-19/Denmark/DCGC-22365/2020\|EPI_ISL_757516\|202 | ···················· | ···················· | ····················· | ···················· | ···················· | ···················· |
| AA | 2726 hCoV-19/Denmark/DCGC-22366/2020\|EPI_ISL_757517\|202 | ···················· | ···················· | ····················· | ···················· | ···················· | ···················· |
| AA | 2727 hCoV-19/Denmark/DCGC-22367/2020\|EPI_ISL_757518\|202 | ···················· | ···················· | ····················· | ···················· | ···················· | ···················· |
| AA | 2728 hCoV-19/Denmark/DCGC-22368/2020\|EPI_ISL_757519\|202 | ···················· | ···················· | ····················· | ···················· | ···················· | ···················· |
| AA | 2729 hCoV-19/Denmark/DCGC-22369/2020\|EPI_ISL_757520\|202 | ···················· | ···················· | ····················· | ···················· | ···················· | ···················· |
| AA | 2730 hCoV-19/Denmark/DCGC-22371/2020\|EPI_ISL_757521\|202 | ···················· | ···················· | ····················· | ···················· | ···················· | ···················· |
| AA | 2731 hCoV-19/Denmark/DCGC-22372/2020\|EPI_ISL_757522\|202 | ···················· | ···················· | ····················· | ···················· | ···················· | ···················· |
| AA | 2732 hCoV-19/Denmark/DCGC-22374/2020\|EPI_ISL_757523\|202 | ···················· | ···················· | ····················· | ···················· | ···················· | ···················· |
| AA | 2733 hCoV-19/Denmark/DCGC-22376/2020\|EPI_ISL_757524\|202 | ···················· | ···················· | ····················· | ···················· | ···················· | ···················· |
| AA | 2734 hCoV-19/Denmark/DCGC-22377/2020\|EPI_ISL_757525\|202 | ···················· | ···················· | ····················· | ···················· | ···················· | ···················· |
| AA | 2735 hCoV-19/Denmark/DCGC-22379/2020\|EPI_ISL_757526\|202 | ···················· | ···················· | ····················· | ···················· | ···················· | ···················· |
| AA | 2736 hCoV-19/Denmark/DCGC-22380/2020\|EPI_ISL_757527\|202 | ···················· | ···················· | ····················· | ···················· | ···················· | ···················· |
| AA | 2737 hCoV-19/Denmark/DCGC-22383/2020\|EPI_ISL_757528\|202 | ···················· | ···················· | ····················· | ···················· | ···················· | ···················· |
| AA | 2738 hCoV-19/Denmark/DCGC-22384/2020\|EPI_ISL_757529\|202 | ···················· | ···················· | ···T················· | ···················· | ···················· | ···················· |
| AA | 2739 hCoV-19/Denmark/DCGC-22385/2020\|EPI_ISL_757530\|202 | ···················· | ···················· | ····················· | ···················· | ···················· | ···················· |
| AA | 2740 hCoV-19/Denmark/DCGC-22386/2020\|EPI_ISL_757531\|202 | ···················· | ···················· | ····················· | ···················· | ···················· | ···················· |
| AA | 2741 hCoV-19/Denmark/DCGC-22388/2020\|EPI_ISL_757532\|202 | ···················· | ···················· | ···A················· | ···················· | ···················· | ···················· |
| AA | 2742 hCoV-19/Denmark/DCGC-22389/2020\|EPI_ISL_757533\|202 | ···················· | ···················· | ····················· | ···················· | ···················· | ···················· |
| AA | 2743 hCoV-19/Denmark/DCGC-22390/2020\|EPI_ISL_757534\|202 | ···················· | ···················· | ····················· | ···················· | ···················· | ···················· |
| AA | 2744 hCoV-19/Denmark/DCGC-22392/2020\|EPI_ISL_757535\|202 | ···················· | ···················· | ····················· | ···················· | ···················· | ···················· |
| AA | 2745 hCoV-19/Denmark/DCGC-22393/2020\|EPI_ISL_757536\|202 | ···················· | ···················· | ····················· | ···················· | ···················· | ···················· |
| AA | 2746 hCoV-19/Denmark/DCGC-22394/2020\|EPI_ISL_757537\|202 | ···················· | ···················· | ····················· | ···················· | ···················· | ···················· |
| AA | 2747 hCoV-19/Denmark/DCGC-22396/2020\|EPI_ISL_757538\|202 | ···················· | ···················· | ····················· | ···················· | ···················· | ···················· |
| AA | 2748 hCoV-19/Denmark/DCGC-22397/2020\|EPI_ISL_757539\|202 | ···················· | ···················· | ····················· | ···················· | ···················· | ···················· |
| AA | 2749 hCoV-19/Denmark/DCGC-22398/2020\|EPI_ISL_757540\|202 | ···················· | ···················· | ····················· | ···················· | ···················· | ···················· |
| AA | 2750 hCoV-19/Denmark/DCGC-22399/2020\|EPI_ISL_757541\|202 | ···················· | ···················· | ····················· | ···················· | ···················· | ···················· |
| AA | 2751 hCoV-19/Denmark/DCGC-22402/2020\|EPI_ISL_757542\|202 | ···················· | ···················· | ····················· | ···················· | ···················· | ···················· |
| AA | 2752 hCoV-19/Denmark/DCGC-22403/2020\|EPI_ISL_757543\|202 | ···················· | ···················· | ····················· | ···················· | ···················· | ···················· |
| AA | 2753 hCoV-19/Denmark/DCGC-22404/2020\|EPI_ISL_757544\|202 | ···················· | ···················· | ····················· | ···················· | ···················· | ···················· |
| AA | 2754 hCoV-19/Denmark/DCGC-22405/2020\|EPI_ISL_757545\|202 | ···················· | ···················· | ····················· | ···················· | ···················· | ···················· |
| AA | 2755 hCoV-19/Denmark/DCGC-22406/2020\|EPI_ISL_757546\|202 | ···················· | ···················· | ····················· | ···················· | ···················· | ···················· |
| AA | 2756 hCoV-19/Denmark/DCGC-22409/2020\|EPI_ISL_757547\|202 | ···················· | ···················· | ····················· | ···················· | ···················· | ···················· |
| AA | 2757 hCoV-19/Denmark/DCGC-22410/2020\|EPI_ISL_757548\|202 | ···················· | ···················· | ····················· | ···················· | ···················· | ···················· |
| AA | 2758 hCoV-19/Denmark/DCGC-22413/2020\|EPI_ISL_757549\|202 | ···················· | ···················· | ····················· | ···················· | ···················· | ···················· |
| AA | 2759 hCoV-19/Denmark/DCGC-22415/2020\|EPI_ISL_757550\|202 | ···················· | ···················· | ····················· | ···················· | ···················· | ···················· |
| AA | 2760 hCoV-19/Denmark/DCGC-22417/2020\|EPI_ISL_757551\|202 | ···················· | ···················· | ····················· | ···················· | ···················· | ···················· |
| AA | 2761 hCoV-19/Denmark/DCGC-22418/2020\|EPI_ISL_757552\|202 | ···················· | ···················· | ····················· | ···················· | ···················· | ···················· |
| AA | 2762 hCoV-19/Denmark/DCGC-22419/2020\|EPI_ISL_757553\|202 | ···················· | ···················· | ····················· | ···················· | ···················· | ···················· |
| AA | 2763 hCoV-19/Denmark/DCGC-22421/2020\|EPI_ISL_757554\|202 | ···················· | ···················· | ····················· | ···················· | ···················· | ···················· |
| AA | 2764 hCoV-19/Denmark/DCGC-22423/2020\|EPI_ISL_757555\|202 | ···················· | ···················· | ····················· | ···················· | ···················· | ···················· |
| AA | 2765 hCoV-19/Denmark/DCGC-22425/2020\|EPI_ISL_757556\|202 | ···················· | ···················· | ····················· | ···················· | ···················· | ···················· |
| AA | 2766 hCoV-19/Denmark/DCGC-22426/2020\|EPI_ISL_757557\|202 | ···················· | ···················· | ····················· | ···················· | ···················· | ···················· |
| AA | 2767 hCoV-19/Denmark/DCGC-22429/2020\|EPI_ISL_757558\|202 | ···················· | ···················· | ····················· | ···················· | ···················· | ···················· |
| AA | 2768 hCoV-19/Denmark/DCGC-22431/2020\|EPI_ISL_757559\|202 | ···················· | ···················· | ····················· | ···················· | ···················· | ···················· |
| AA | 2769 hCoV-19/Denmark/DCGC-22433/2020\|EPI_ISL_757560\|202 | ···················· | ···················· | ····················· | ···················· | ···················· | ···················· |
| AA | 2770 hCoV-19/Denmark/DCGC-22437/2020\|EPI_ISL_757561\|202 | ···················· | ···················· | ····················· | ···················· | ···················· | ···················· |
| AA | 2771 hCoV-19/Denmark/DCGC-22438/2020\|EPI_ISL_757562\|202 | ···················· | ···················· | ····················· | ···················· | ···················· | ···················· |
| AA | 2772 hCoV-19/Denmark/DCGC-22439/2020\|EPI_ISL_757563\|202 | ···················· | ···················· | ····················· | ···················· | ···················· | ···················· |
| AA | 2773 hCoV-19/Denmark/DCGC-22440/2020\|EPI_ISL_757564\|202 | ···················· | ···················· | ····················· | ···················· | ···················· | ···················· |
| AA | 2774 hCoV-19/Denmark/DCGC-22442/2020\|EPI_ISL_757565\|202 | ···················· | ···················· | ····················· | ···················· | ···················· | ···················· |
| AA | 2775 hCoV-19/Denmark/DCGC-22443/2020\|EPI_ISL_757566\|202 | ···················· | ···················· | ····················· | ···················· | ···················· | ···················· |
| AA | 2776 hCoV-19/Denmark/DCGC-22445/2020\|EPI_ISL_757567\|202 | ···················· | ···················· | ····················· | ···················· | ···················· | ···················· |
| AA | 2777 hCoV-19/Denmark/DCGC-22448/2020\|EPI_ISL_757568\|202 | ···················· | ···················· | ····················· | ···················· | ···················· | ···················· |
| AA | 2778 hCoV-19/Denmark/DCGC-22449/2020\|EPI_ISL_757569\|202 | ···················· | ···················· | ····················· | ···················· | ···················· | ···················· |
| AA | 2779 hCoV-19/Denmark/DCGC-22450/2020\|EPI_ISL_757570\|202 | ···················· | ···················· | ····················· | ···················· | ···················· | ···················· |
| AA | 2780 hCoV-19/Denmark/DCGC-22453/2020\|EPI_ISL_757571\|202 | ···················· | ···················· | ····················· | ···················· | ···················· | ···················· |
| AA | 2781 hCoV-19/Denmark/DCGC-22457/2020\|EPI_ISL_757572\|202 | ···················· | ···················· | ····················· | ···················· | ···················· | ···················· |
| AA | 2782 hCoV-19/Denmark/DCGC-22461/2020\|EPI_ISL_757573\|202 | ···················· | ···················· | ····················· | ···················· | ···················· | ···················· |
| AA | 2783 hCoV-19/Denmark/DCGC-22463/2020\|EPI_ISL_757574\|202 | ···················· | ···················· | ····················· | ···················· | ···················· | ···················· |
| AA | 2784 hCoV-19/Denmark/DCGC-22466/2020\|EPI_ISL_757575\|202 | ···················· | ···················· | ····················· | ···················· | ···················· | ···················· |
| AA | 2785 hCoV-19/Denmark/DCGC-22467/2020\|EPI_ISL_757576\|202 | ···················· | ···················· | ····················· | ···················· | ···················· | ···················· |
| AA | 2786 hCoV-19/Denmark/DCGC-22468/2020\|EPI_ISL_757577\|202 | ···················· | ···················· | ····················· | ···················· | ···················· | ···················· |
| AA | 2787 hCoV-19/Denmark/DCGC-22469/2020\|EPI_ISL_757578\|202 | ···················· | ···················· | ····················· | ···················· | ···················· | ···················· |
| AA | 2788 hCoV-19/Denmark/DCGC-22471/2020\|EPI_ISL_757579\|202 | ···················· | ·A·················· | ····················· | ···················· | ···················· | ···A················ |
| AA | 2789 hCoV-19/Denmark/DCGC-22472/2020\|EPI_ISL_757580\|202 | ···················· | ···················· | ····················· | ···················· | ···················· | ···················· |
| AA | 2790 hCoV-19/Denmark/DCGC-22474/2020\|EPI_ISL_757581\|202 | ···················· | ···················· | ····················· | ···················· | ···················· | ···················· |
| AA | 2791 hCoV-19/Denmark/DCGC-22475/2020\|EPI_ISL_757582\|202 | ···················· | ···················· | ····················· | ···················· | ···················· | ···················· |
| AA | 2792 hCoV-19/Denmark/DCGC-22476/2020\|EPI_ISL_757583\|202 | ···················· | ···················· | ····················· | ···················· | ···················· | ···················· |
| AA | 2793 hCoV-19/Denmark/DCGC-22477/2020\|EPI_ISL_757584\|202 | ···················· | ···················· | ····················· | ···················· | ···················· | ···················· |
| AA | 2794 hCoV-19/Denmark/DCGC-22480/2020\|EPI_ISL_757585\|202 | ···················· | ···················· | ····················· | ···················· | ···················· | ···················· |
| AA | 2795 hCoV-19/Denmark/DCGC-22484/2020\|EPI_ISL_757586\|202 | ···················· | ···················· | ····················· | ···················· | ···················· | ···················· |
| AA | 2796 hCoV-19/Denmark/DCGC-22485/2020\|EPI_ISL_757587\|202 | ···················· | ···················· | ····················· | ···················· | ···················· | ···················· |
| AA | 2797 hCoV-19/Denmark/DCGC-22486/2020\|EPI_ISL_757588\|202 | ···················· | ···················· | ····················· | ···················· | ···················· | ···················· |
| AA | 2798 hCoV-19/Denmark/DCGC-22488/2020\|EPI_ISL_757589\|202 | ···················· | ···················· | ····················· | ···················· | ···················· | ···················· |
| AA | 2799 hCoV-19/Denmark/DCGC-22489/2020\|EPI_ISL_757590\|202 | ···················· | ···················· | ····················· | ···················· | ···················· | ···················· |
| AA | 2800 hCoV-19/Denmark/DCGC-22490/2020\|EPI_ISL_757591\|202 | ···················· | ···················· | ····················· | ···················· | ···················· | ···················· |
| AA | 2801 hCoV-19/Denmark/DCGC-22493/2020\|EPI_ISL_757592\|202 | ···················· | ···················· | ····················· | ···················· | ···················· | ···················· |
| AA | 2802 hCoV-19/Denmark/DCGC-22495/2020\|EPI_ISL_757593\|202 | ···················· | ···················· | ····················· | ···················· | ···················· | ···················· |
| AA | 2803 hCoV-19/Denmark/DCGC-22498/2020\|EPI_ISL_757594\|202 | ···················· | ···················· | ····················· | ···················· | ···················· | ···················· |

| | | | | | | | | |
|---|---|---|---|---|---|---|---|---|
| AA | 2804 | hCoV-19/Denmark/DCGC-22501/2020\|EPI_ISL_757595\|202 | ···················· | ···················· | ····················· | ···················· | ···················· | ···················· |
| AA | 2805 | hCoV-19/Denmark/DCGC-22502/2020\|EPI_ISL_757596\|202 | ···················· | ···················· | ····················· | ···················· | ···················· | ···················· |
| AA | 2806 | hCoV-19/Denmark/DCGC-22503/2020\|EPI_ISL_757597\|202 | ···················· | ···················· | ····················· | ···················· | ···················· | ···················· |
| AA | 2807 | hCoV-19/Denmark/DCGC-22504/2020\|EPI_ISL_757598\|202 | ···················· | ···················· | ····················· | ···················· | ···················· | ···················· |
| AA | 2808 | hCoV-19/Denmark/DCGC-22505/2020\|EPI_ISL_757599\|202 | ···················· | ···················· | ····················· | ···················· | ···················· | ···················· |
| AA | 2809 | hCoV-19/Denmark/DCGC-20360/2020\|EPI_ISL_758892\|202 | ···················· | ···················· | ····················· | ···················· | ···················· | ···················· |
| AA | 2810 | hCoV-19/Denmark/DCGC-20364/2020\|EPI_ISL_758893\|202 | ···················· | ···················· | ····················· | ···················· | ···················· | ···················· |
| AA | 2811 | hCoV-19/Denmark/DCGC-20536/2020\|EPI_ISL_758894\|202 | ···················· | ···················· | ····················· | ···················· | ···················· | ···················· |
| AA | 2812 | hCoV-19/Denmark/DCGC-20881/2020\|EPI_ISL_758895\|202 | ···················· | ···················· | ····················· | ···················· | ···················· | ···················· |
| AA | 2813 | hCoV-19/Denmark/DCGC-21082/2020\|EPI_ISL_758896\|202 | ···················· | ···················· | ····················· | ···················· | ···················· | ···················· |
| AA | 2814 | hCoV-19/Denmark/DCGC-20400/2020\|EPI_ISL_758897\|202 | ···················· | ···················· | ····················· | ···················· | ···················· | ···················· |
| AA | 2815 | hCoV-19/Denmark/DCGC-20458/2020\|EPI_ISL_758898\|202 | ···················· | ···················· | ····················· | ···················· | ···················· | ···················· |
| AA | 2816 | hCoV-19/Denmark/DCGC-20515/2020\|EPI_ISL_758899\|202 | ···················· | ···················· | ····················· | ···················· | ···················· | ···················· |
| AA | 2817 | hCoV-19/Denmark/DCGC-19670/2020\|EPI_ISL_749100\|202 | ···················· | ···················· | ····················· | ···················· | ···················· | ···················· |
| AA | 2818 | hCoV-19/Denmark/DCGC-19671/2020\|EPI_ISL_749101\|202 | ···················· | ···················· | ····················· | ···················· | ···················· | ···················· |
| AA | 2819 | hCoV-19/Denmark/DCGC-19672/2020\|EPI_ISL_749102\|202 | ···················· | ···················· | ····················· | ···················· | ···················· | ···················· |
| AA | 2820 | hCoV-19/Denmark/DCGC-19673/2020\|EPI_ISL_749103\|202 | ···················· | ···················· | ····················· | ···················· | ···················· | ···················· |
| AA | 2821 | hCoV-19/Denmark/DCGC-19675/2020\|EPI_ISL_749104\|202 | ···················· | ···················· | ····················· | ···················· | ···················· | ···················· |
| AA | 2822 | hCoV-19/Denmark/DCGC-19676/2020\|EPI_ISL_749105\|202 | ···················· | ···················· | ····················· | ···················· | ···················· | ···················· |
| AA | 2823 | hCoV-19/Denmark/DCGC-19679/2020\|EPI_ISL_749106\|202 | ···················· | ·A·················· | ····················· | ···················· | ···················· | ···················· |
| AA | 2824 | hCoV-19/Denmark/DCGC-19680/2020\|EPI_ISL_749107\|202 | ···················· | ···················· | ····················· | ···················· | ···················· | ···················· |
| AA | 2825 | hCoV-19/Denmark/DCGC-19682/2020\|EPI_ISL_749108\|202 | ···················· | ···················· | ····················· | ···················· | ···················· | ···················· |
| AA | 2826 | hCoV-19/Denmark/DCGC-19685/2020\|EPI_ISL_749109\|202 | ···················· | ···················· | ····················· | ···················· | ···················· | ···················· |
| AA | 2827 | hCoV-19/Denmark/DCGC-19686/2020\|EPI_ISL_749110\|202 | ···················· | ···················· | ····················· | ···················· | ···················· | ···················· |
| AA | 2828 | hCoV-19/Denmark/DCGC-19689/2020\|EPI_ISL_749111\|202 | ···················· | ···················· | ····················· | ···················· | ···················· | ···················· |
| AA | 2829 | hCoV-19/Denmark/DCGC-19690/2020\|EPI_ISL_749112\|202 | ···················· | ···················· | ····················· | ···················· | ···················· | ···················· |
| AA | 2830 | hCoV-19/Denmark/DCGC-19691/2020\|EPI_ISL_749113\|202 | ···················· | ···················· | ····················· | ···················· | ···················· | ···················· |
| AA | 2831 | hCoV-19/Denmark/DCGC-19692/2020\|EPI_ISL_749114\|202 | ···················· | ···················· | ····················· | ···················· | ···················· | ···················· |
| AA | 2832 | hCoV-19/Denmark/DCGC-19693/2020\|EPI_ISL_749115\|202 | ···················· | ···················· | ····················· | ···················· | ···················· | ···················· |
| AA | 2833 | hCoV-19/Denmark/DCGC-19694/2020\|EPI_ISL_749116\|202 | ···················· | ···················· | ····················· | ···················· | ···················· | ···················· |
| AA | 2834 | hCoV-19/Denmark/DCGC-19695/2020\|EPI_ISL_749117\|202 | ···················· | ·A·················· | ····················· | ···················· | ···················· | ··A················· |
| AA | 2835 | hCoV-19/Denmark/DCGC-19698/2020\|EPI_ISL_749118\|202 | ···················· | ···················· | ····················· | ···················· | ···················· | ···················· |
| AA | 2836 | hCoV-19/Denmark/DCGC-19699/2020\|EPI_ISL_749119\|202 | ···················· | ···················· | ····················· | ···················· | ···················· | ···················· |
| AA | 2837 | hCoV-19/Denmark/DCGC-19700/2020\|EPI_ISL_749120\|202 | ···················· | ···················· | ····················· | ···················· | ···················· | ···················· |
| AA | 2838 | hCoV-19/Denmark/DCGC-19701/2020\|EPI_ISL_749121\|202 | ···················· | ···················· | ····················· | ···················· | ···················· | ···················· |
| AA | 2839 | hCoV-19/Denmark/DCGC-19705/2020\|EPI_ISL_749122\|202 | ···················· | ···················· | ····················· | ···················· | ···················· | ···················· |
| AA | 2840 | hCoV-19/Denmark/DCGC-19706/2020\|EPI_ISL_749123\|202 | ···················· | ···················· | ····················· | ···················· | ···················· | ···················· |
| AA | 2841 | hCoV-19/Denmark/DCGC-19707/2020\|EPI_ISL_749124\|202 | ···················· | ···················· | ····················· | ···················· | ···················· | ···················· |
| AA | 2842 | hCoV-19/Denmark/DCGC-19710/2020\|EPI_ISL_749125\|202 | ···················· | ···················· | ····················· | ···················· | ···················· | ···················· |
| AA | 2843 | hCoV-19/Denmark/DCGC-19711/2020\|EPI_ISL_749126\|202 | ···················· | ···················· | ····················· | ···················· | ···················· | ···················· |
| AA | 2844 | hCoV-19/Denmark/DCGC-19712/2020\|EPI_ISL_749127\|202 | ···················· | ···················· | ····················· | ···················· | ···················· | ···················· |
| AA | 2845 | hCoV-19/Denmark/DCGC-19714/2020\|EPI_ISL_749128\|202 | ···················· | ···················· | ····················· | ···················· | ···················· | ···················· |
| AA | 2846 | hCoV-19/Denmark/DCGC-19715/2020\|EPI_ISL_749129\|202 | ···················· | ···················· | ····················· | ···················· | ···················· | ···················· |
| AA | 2847 | hCoV-19/Denmark/DCGC-19716/2020\|EPI_ISL_749130\|202 | ···················· | ···················· | ····················· | ···················· | ···················· | ···················· |
| AA | 2848 | hCoV-19/Denmark/DCGC-19717/2020\|EPI_ISL_749131\|202 | ···················· | ···················· | ····················· | ···················· | ···················· | ···················· |
| AA | 2849 | hCoV-19/Denmark/DCGC-19719/2020\|EPI_ISL_749132\|202 | ···················· | ···················· | ····················· | ···················· | ···················· | ···················· |
| AA | 2850 | hCoV-19/Denmark/DCGC-19720/2020\|EPI_ISL_749133\|202 | ···················· | ···················· | ····················· | ···················· | ···················· | ···················· |
| AA | 2851 | hCoV-19/Denmark/DCGC-19722/2020\|EPI_ISL_749134\|202 | ···················· | ···················· | ····················· | ···················· | ···················· | ···················· |
| AA | 2852 | hCoV-19/Denmark/DCGC-19723/2020\|EPI_ISL_749135\|202 | ···················· | ···················· | ····················· | ···················· | ···················· | ···················· |
| AA | 2853 | hCoV-19/Denmark/DCGC-19726/2020\|EPI_ISL_749136\|202 | ···················· | ···················· | ····················· | ···················· | ···················· | ···················· |
| AA | 2854 | hCoV-19/Denmark/DCGC-19727/2020\|EPI_ISL_749137\|202 | ···················· | ·A·················· | ····················· | ···················· | ···················· | ··A················· |
| AA | 2855 | hCoV-19/Denmark/DCGC-19729/2020\|EPI_ISL_749138\|202 | ···················· | ···················· | ····················· | ···················· | ···················· | ···················· |
| AA | 2856 | hCoV-19/Denmark/DCGC-19730/2020\|EPI_ISL_749139\|202 | ···················· | ···················· | ····················· | ···················· | ···················· | ···················· |
| AA | 2857 | hCoV-19/Denmark/DCGC-19731/2020\|EPI_ISL_749140\|202 | ···················· | ···················· | ····················· | ···················· | ···················· | ···················· |
| AA | 2858 | hCoV-19/Denmark/DCGC-19736/2020\|EPI_ISL_749141\|202 | ···················· | ···················· | ····················· | ···················· | ···················· | ···················· |
| AA | 2859 | hCoV-19/Denmark/DCGC-19737/2020\|EPI_ISL_749142\|202 | ···················· | ···················· | ····················· | ···················· | ···················· | ···················· |
| AA | 2860 | hCoV-19/Denmark/DCGC-19738/2020\|EPI_ISL_749143\|202 | ···················· | ·A·················· | ····················· | ···················· | ···················· | ···················· |
| AA | 2861 | hCoV-19/Denmark/DCGC-19739/2020\|EPI_ISL_749144\|202 | ···················· | ···················· | ····················· | ···················· | ···················· | ···················· |
| AA | 2862 | hCoV-19/Denmark/DCGC-19743/2020\|EPI_ISL_749145\|202 | ···················· | ···················· | ····················· | ···················· | ···················· | ···················· |
| AA | 2863 | hCoV-19/Denmark/DCGC-19744/2020\|EPI_ISL_749146\|202 | ···················· | ···················· | ····················· | ···················· | ···················· | ···················· |
| AA | 2864 | hCoV-19/Denmark/DCGC-19746/2020\|EPI_ISL_749147\|202 | ···················· | ···················· | ····················· | ···················· | ···················· | ···················· |
| AA | 2865 | hCoV-19/Uruguay/TYT-M49/2020\|EPI_ISL_749148\|2020-0 | ···················· | ···················· | ····················· | ···················· | ···················· | ···················· |
| AA | 2866 | hCoV-19/Uruguay/TYT-M57/2020\|EPI_ISL_749151\|2020-0 | ···················· | ···················· | ····················· | ···················· | ···················· | ···················· |
| AA | 2867 | hCoV-19/Uruguay/TYT-M64/2020\|EPI_ISL_749155\|2020-0 | ···················· | ···················· | ····················· | ···················· | ···················· | ···················· |
| AA | 2868 | hCoV-19/Denmark/DCGC-18647/2020\|EPI_ISL_750100\|202 | ···················· | ···················· | ····················· | ···················· | ···················· | ···················· |
| AA | 2869 | hCoV-19/Denmark/DCGC-18648/2020\|EPI_ISL_750101\|202 | ···················· | ···················· | ····················· | ···················· | ···················· | ···················· |
| AA | 2870 | hCoV-19/Denmark/DCGC-18649/2020\|EPI_ISL_750102\|202 | ···················· | ···················· | ····················· | ···················· | ···················· | ···················· |
| AA | 2871 | hCoV-19/Denmark/DCGC-18653/2020\|EPI_ISL_750103\|202 | ···················· | ···················· | ····················· | ···················· | ···················· | ···················· |
| AA | 2872 | hCoV-19/Denmark/DCGC-18656/2020\|EPI_ISL_750104\|202 | ···················· | ···················· | ····················· | ···················· | ···················· | ···················· |
| AA | 2873 | hCoV-19/Denmark/DCGC-18657/2020\|EPI_ISL_750105\|202 | ···················· | ·A·················· | ····················· | ···················· | ···················· | ··A················· |
| AA | 2874 | hCoV-19/Denmark/DCGC-18659/2020\|EPI_ISL_750106\|202 | ···················· | ···················· | ····················· | ···················· | ···················· | ···················· |
| AA | 2875 | hCoV-19/Denmark/DCGC-18660/2020\|EPI_ISL_750107\|202 | ···················· | ···················· | ····················· | ···················· | ···················· | ···················· |
| AA | 2876 | hCoV-19/Denmark/DCGC-18661/2020\|EPI_ISL_750109\|202 | ···················· | ···················· | ····················· | ···················· | ···················· | ···················· |
| AA | 2877 | hCoV-19/Denmark/DCGC-18662/2020\|EPI_ISL_750110\|202 | ···················· | ···················· | ····················· | ···················· | ···················· | ···················· |
| AA | 2878 | hCoV-19/Denmark/DCGC-18663/2020\|EPI_ISL_750111\|202 | ···················· | ···················· | ····················· | ···················· | ···················· | ···················· |
| AA | 2879 | hCoV-19/Denmark/DCGC-18664/2020\|EPI_ISL_750112\|202 | ···················· | ···················· | ····················· | ···················· | ···················· | ···················· |
| AA | 2880 | hCoV-19/Denmark/DCGC-18666/2020\|EPI_ISL_750113\|202 | ···················· | ···················· | ····················· | ···················· | ···················· | ···················· |
| AA | 2881 | hCoV-19/Denmark/DCGC-18669/2020\|EPI_ISL_750114\|202 | ···················· | ···················· | ····················· | ···················· | ···················· | ···················· |
| AA | 2882 | hCoV-19/Denmark/DCGC-18670/2020\|EPI_ISL_750115\|202 | ···················· | ···················· | ····················· | ···················· | ···················· | ···················· |
| AA | 2883 | hCoV-19/Denmark/DCGC-18671/2020\|EPI_ISL_750116\|202 | ···················· | ···················· | ····················· | ···················· | ···················· | ···················· |

| | | | | | | | | |
|---|---|---|---|---|---|---|---|---|
| AA | 2884 | hCoV-19/Denmark/DCGC-18674/2020\|EPI_ISL_750117\|202 | ···················· | ···················· | ····················· | ···················· | ···················· | ···················· |
| AA | 2885 | hCoV-19/Denmark/DCGC-18675/2020\|EPI_ISL_750118\|202 | ···················· | ···················· | ····················· | ···················· | ···················· | ···················· |
| AA | 2886 | hCoV-19/Denmark/DCGC-18676/2020\|EPI_ISL_750119\|202 | ···················· | ···················· | ····················· | ···················· | ···················· | ···················· |
| AA | 2887 | hCoV-19/Denmark/DCGC-18677/2020\|EPI_ISL_750120\|202 | ···················· | ···················· | ····················· | ···················· | ···················· | ···················· |
| AA | 2888 | hCoV-19/Denmark/DCGC-18678/2020\|EPI_ISL_750121\|202 | ···················· | ···················· | ····················· | ···················· | ···················· | ···················· |
| AA | 2889 | hCoV-19/Denmark/DCGC-18680/2020\|EPI_ISL_750122\|202 | ···················· | ···················· | ··A·················· | ···················· | ···················· | ···················· |
| AA | 2890 | hCoV-19/Denmark/DCGC-18685/2020\|EPI_ISL_750123\|202 | ···················· | ···················· | ····················· | ···················· | ···················· | ···················· |
| AA | 2891 | hCoV-19/Denmark/DCGC-18686/2020\|EPI_ISL_750124\|202 | ···················· | ···················· | ····················· | ···················· | ···················· | ···················· |
| AA | 2892 | hCoV-19/Denmark/DCGC-18687/2020\|EPI_ISL_750125\|202 | ···················· | ···················· | ····················· | ···················· | ···················· | ···················· |
| AA | 2893 | hCoV-19/Denmark/DCGC-18689/2020\|EPI_ISL_750126\|202 | ···················· | ···················· | ····················· | ···················· | ···················· | ···················· |
| AA | 2894 | hCoV-19/Denmark/DCGC-18692/2020\|EPI_ISL_750127\|202 | ···················· | ···················· | ····················· | ···················· | ···················· | ···················· |
| AA | 2895 | hCoV-19/Denmark/DCGC-18694/2020\|EPI_ISL_750128\|202 | ···················· | ···················· | ····················· | ···················· | ···················· | ···················· |
| AA | 2896 | hCoV-19/Denmark/DCGC-18695/2020\|EPI_ISL_750129\|202 | ···················· | ···················· | ····················· | ···················· | ···················· | ···················· |
| AA | 2897 | hCoV-19/Denmark/DCGC-18697/2020\|EPI_ISL_750130\|202 | ···················· | ···················· | ····················· | ···················· | ···················· | ···················· |
| AA | 2898 | hCoV-19/Denmark/DCGC-18701/2020\|EPI_ISL_750132\|202 | ···················· | ·A·················· | ····················· | ···················· | ···················· | ··A················· |
| AA | 2899 | hCoV-19/Denmark/DCGC-18704/2020\|EPI_ISL_750133\|202 | ···················· | ···················· | ····················· | ···················· | ···················· | ···················· |
| AA | 2900 | hCoV-19/Denmark/DCGC-18705/2020\|EPI_ISL_750134\|202 | ···················· | ···················· | ····················· | ···················· | ···················· | ···················· |
| AA | 2901 | hCoV-19/Denmark/DCGC-18709/2020\|EPI_ISL_750135\|202 | ···················· | ·A·················· | ····················· | ···················· | ···················· | ···················· |
| AA | 2902 | hCoV-19/Denmark/DCGC-18726/2020\|EPI_ISL_750147\|202 | ···················· | ···················· | ····················· | ···················· | ···················· | ···················· |
| AA | 2903 | hCoV-19/Denmark/DCGC-18728/2020\|EPI_ISL_750148\|202 | ···················· | ···················· | ····················· | ···················· | ···················· | ···················· |
| AA | 2904 | hCoV-19/Denmark/DCGC-18729/2020\|EPI_ISL_750149\|202 | ···················· | ···················· | ····················· | ···················· | ···················· | ···················· |
| AA | 2905 | hCoV-19/Denmark/DCGC-18732/2020\|EPI_ISL_750150\|202 | ···················· | ···················· | ····················· | ···················· | ···················· | ···················· |
| AA | 2906 | hCoV-19/Denmark/DCGC-18733/2020\|EPI_ISL_750151\|202 | ···················· | ···················· | ····················· | ···················· | ···················· | ···················· |
| AA | 2907 | hCoV-19/Denmark/DCGC-18734/2020\|EPI_ISL_750152\|202 | ···················· | ···················· | ····················· | ···················· | ···················· | ···················· |
| AA | 2908 | hCoV-19/Denmark/DCGC-18736/2020\|EPI_ISL_750153\|202 | ···················· | ···················· | ····················· | ···················· | ···················· | ···················· |
| AA | 2909 | hCoV-19/Denmark/DCGC-18737/2020\|EPI_ISL_750154\|202 | ···················· | ···················· | ····················· | ···················· | ···················· | ···················· |
| AA | 2910 | hCoV-19/Denmark/DCGC-18739/2020\|EPI_ISL_750155\|202 | ···················· | ···················· | ····················· | ···················· | ···················· | ···················· |
| AA | 2911 | hCoV-19/Denmark/DCGC-18741/2020\|EPI_ISL_750156\|202 | ···················· | ···················· | ····················· | ···················· | ···················· | ···················· |
| AA | 2912 | hCoV-19/Denmark/DCGC-18742/2020\|EPI_ISL_750157\|202 | ···················· | ···················· | ····················· | ···················· | ···················· | ···················· |
| AA | 2913 | hCoV-19/Denmark/DCGC-18744/2020\|EPI_ISL_750158\|202 | ···················· | ···················· | ····················· | ···················· | ···················· | ···················· |
| AA | 2914 | hCoV-19/Denmark/DCGC-18745/2020\|EPI_ISL_750159\|202 | ···················· | ···················· | ····················· | ···················· | ···················· | ···················· |
| AA | 2915 | hCoV-19/Denmark/DCGC-18746/2020\|EPI_ISL_750160\|202 | ···················· | ···················· | ····················· | ···················· | ···················· | ···················· |
| AA | 2916 | hCoV-19/Uruguay/TYT-M87/2020\|EPI_ISL_750168\|2020-0 | ···················· | ···················· | ····················· | ···················· | ···················· | ···················· |
| AA | 2917 | hCoV-19/Uruguay/TYT-M91/2020\|EPI_ISL_750169\|2020-0 | ···················· | ···················· | ····················· | ···················· | ···················· | ···················· |
| AA | 2918 | hCoV-19/Uruguay/TYT-M93/2020\|EPI_ISL_750170\|2020-0 | ···················· | ···················· | ····················· | ···················· | ···················· | ···················· |
| AA | 2919 | hCoV-19/Uruguay/TYT-M97/2020\|EPI_ISL_750173\|2020-0 | ···················· | ···················· | ····················· | ···················· | ···················· | ···················· |
| AA | 2920 | hCoV-19/Uruguay/CEL-M116/2020\|EPI_ISL_750177\|2020- | ···················· | ···················· | ····················· | ···················· | ···················· | ···················· |
| AA | 2921 | hCoV-19/Uruguay/RIV-M119/2020\|EPI_ISL_750178\|2020- | ···················· | ···················· | ····················· | ···················· | ···················· | ···················· |
| AA | 2922 | hCoV-19/Italy/VEN-UniVR-108/2020\|EPI_ISL_751412\|20 | ···················· | ···················· | ····················· | ···················· | ···················· | ···················· |
| AA | 2923 | hCoV-19/Italy/VEN-UniVR-109/2020\|EPI_ISL_751413\|20 | ···················· | ···················· | ····················· | ···················· | ···················· | ···················· |
| AA | 2924 | hCoV-19/Italy/VEN-UniVR-111/2020\|EPI_ISL_751414\|20 | ···················· | ···················· | ····················· | ···················· | ···················· | ···················· |
| AA | 2925 | hCoV-19/Italy/VEN-UniVR-112/2020\|EPI_ISL_751415\|20 | ···················· | ···················· | ····················· | ···················· | ···················· | ···················· |
| AA | 2926 | hCoV-19/Italy/VEN-UniVR-113/2020\|EPI_ISL_751416\|20 | ···················· | ···················· | ····················· | ···················· | ···················· | ···················· |
| AA | 2927 | hCoV-19/Italy/VEN-UniVR-116/2020\|EPI_ISL_751417\|20 | ···················· | ···················· | ····················· | ···················· | ···················· | ···················· |
| AA | 2928 | hCoV-19/Italy/VEN-UniVR-120/2020\|EPI_ISL_751420\|20 | ···················· | ···················· | ····················· | ···················· | ···················· | ···················· |
| AA | 2929 | hCoV-19/Italy/VEN-UniVR-121/2020\|EPI_ISL_751421\|20 | ···················· | ···················· | ····················· | ···················· | ···················· | ···················· |
| AA | 2930 | hCoV-19/Italy/VEN-UniVR-122/2020\|EPI_ISL_751422\|20 | ···················· | ···················· | ····················· | ···················· | ···················· | ···················· |
| AA | 2931 | hCoV-19/Italy/VEN-UniVR-123/2020\|EPI_ISL_751423\|20 | ···················· | ···················· | ····················· | ···················· | ···················· | ···················· |
| AA | 2932 | hCoV-19/Italy/VEN-UniVR-124/2020\|EPI_ISL_751424\|20 | ···················· | ···················· | ····················· | ···················· | ···················· | ···················· |
| AA | 2933 | hCoV-19/Italy/VEN-UniVR-125/2020\|EPI_ISL_751425\|20 | ···················· | ···················· | ····················· | ···················· | ···················· | ···················· |
| AA | 2934 | hCoV-19/Italy/VEN-UniVR-126/2020\|EPI_ISL_751426\|20 | ···················· | ···················· | ····················· | ···················· | ···················· | ···················· |
| AA | 2935 | hCoV-19/Italy/VEN-UniVR-128/2020\|EPI_ISL_751427\|20 | ···················· | ···················· | ····················· | ···················· | ···················· | ···················· |
| AA | 2936 | hCoV-19/Italy/VEN-UniVR-129/2020\|EPI_ISL_751428\|20 | ···················· | ···················· | ····················· | ···················· | ···················· | ···················· |
| AA | 2937 | hCoV-19/Italy/VEN-UniVR-132/2020\|EPI_ISL_751429\|20 | ···················· | ···················· | ····················· | ···················· | ···················· | ···················· |
| AA | 2938 | hCoV-19/Italy/VEN-UniVR-133/2020\|EPI_ISL_751430\|20 | ···················· | ···················· | ····················· | ···················· | ···················· | ···················· |
| AA | 2939 | hCoV-19/Italy/VEN-UniVR-134/2020\|EPI_ISL_751431\|20 | ···················· | ···················· | ····················· | ···················· | ···················· | ···················· |
| AA | 2940 | hCoV-19/Italy/VEN-UniVR-135/2020\|EPI_ISL_751432\|20 | ···················· | ···················· | ····················· | ···················· | ···················· | ···················· |
| AA | 2941 | hCoV-19/Italy/VEN-UniVR-136/2020\|EPI_ISL_751433\|20 | ···················· | ···················· | ····················· | ···················· | ···················· | ···················· |
| AA | 2942 | hCoV-19/Italy/VEN-UniVR-137/2020\|EPI_ISL_751434\|20 | ···················· | ···················· | ····················· | ···················· | ···················· | ···················· |
| AA | 2943 | hCoV-19/Italy/VEN-UniVR-138/2020\|EPI_ISL_751435\|20 | ···················· | ···················· | ····················· | ···················· | ···················· | ···················· |
| AA | 2944 | hCoV-19/Italy/VEN-UniVR-139/2020\|EPI_ISL_751436\|20 | ···················· | ···················· | ····················· | ···················· | ···················· | ···················· |
| AA | 2945 | hCoV-19/Italy/VEN-UniVR-140/2020\|EPI_ISL_751437\|20 | ···················· | ···················· | ····················· | ···················· | ···················· | ···················· |
| AA | 2946 | hCoV-19/Italy/VEN-UniVR-141/2020\|EPI_ISL_751438\|20 | ···················· | ···················· | ····················· | ···················· | ···················· | ···················· |
| AA | 2947 | hCoV-19/Italy/VEN-UniVR-142/2020\|EPI_ISL_751439\|20 | ···················· | ···················· | ····················· | ···················· | ···················· | ···················· |
| AA | 2948 | hCoV-19/Italy/VEN-UniVR-143/2020\|EPI_ISL_751440\|20 | ···················· | ···················· | ····················· | ···················· | ···················· | ···················· |
| AA | 2949 | hCoV-19/Italy/VEN-UniVR-144/2020\|EPI_ISL_751441\|20 | ···················· | ···················· | ····················· | ···················· | ···················· | ···················· |
| AA | 2950 | hCoV-19/Italy/VEN-UniVR-145/2020\|EPI_ISL_751442\|20 | ···················· | ···················· | ····················· | ···················· | ···················· | ···················· |
| AA | 2951 | hCoV-19/Italy/VEN-UniVR-146/2020\|EPI_ISL_751443\|20 | ···················· | ···················· | ····················· | ···················· | ···················· | ···················· |
| AA | 2952 | hCoV-19/Italy/VEN-UniVR-147/2020\|EPI_ISL_751444\|20 | ···················· | ···················· | ····················· | ···················· | ···················· | ···················· |
| AA | 2953 | hCoV-19/Italy/VEN-UniVR-150/2020\|EPI_ISL_751445\|20 | ···················· | ···················· | ····················· | ···················· | ···················· | ···················· |
| AA | 2954 | hCoV-19/Italy/VEN-UniVR-152/2020\|EPI_ISL_751446\|20 | ···················· | ···················· | ····················· | ···················· | ···················· | ···················· |
| AA | 2955 | hCoV-19/Italy/VEN-UniVR-153/2020\|EPI_ISL_751447\|20 | ···················· | ···················· | ····················· | ···················· | ···················· | ···················· |
| AA | 2956 | hCoV-19/France/OCC-128/2020\|EPI_ISL_751448\|2020-10 | ···················· | ···················· | ····················· | ···················· | ···················· | ···················· |
| AA | 2957 | hCoV-19/France/OCC-130/2020\|EPI_ISL_751450\|2020-11 | ···················· | ···················· | ····················· | ···················· | ···················· | ···················· |
| AA | 2958 | hCoV-19/France/OCC-131/2020\|EPI_ISL_751451\|2020-11 | ···················· | ···················· | ····················· | ···················· | ···················· | ···················· |
| AA | 2959 | hCoV-19/France/OCC-132/2020\|EPI_ISL_751452\|2020-11 | ···················· | ···················· | ····················· | ···················· | ···················· | ···················· |
| AA | 2960 | hCoV-19/France/OCC-133/2020\|EPI_ISL_751453\|2020-11 | ···················· | ···················· | ····················· | ···················· | ···················· | ···················· |
| AA | 2961 | hCoV-19/France/OCC-134/2020\|EPI_ISL_751454\|2020-11 | ···················· | ···················· | ····················· | ···················· | ···················· | ···················· |
| AA | 2962 | hCoV-19/France/OCC-135/2020\|EPI_ISL_751455\|2020-11 | ···················· | ···················· | ····················· | ···················· | ···················· | ···················· |
| AA | 2963 | hCoV-19/France/OCC-136/2020\|EPI_ISL_751456\|2020-11 | ···················· | ···················· | ····················· | ···················· | ···················· | ···················· |

| | | | | | | | | |
|---|---|---|---|---|---|---|---|---|
| AA | 2964 | hCoV-19/France/OCC-137/2020\|EPI_ISL_751457\|2020-11 | ···················· | ···················· | ····················· | ···················· | ···················· | ···················· |
| AA | 2965 | hCoV-19/France/OCC-138/2020\|EPI_ISL_751458\|2020-11 | ···················· | ···················· | ····················· | ···················· | ···················· | ···················· |
| AA | 2966 | hCoV-19/France/OCC-141/2020\|EPI_ISL_751461\|2020-11 | ···················· | ···················· | ····················· | ···················· | ···················· | ···················· |
| AA | 2967 | hCoV-19/France/OCC-143/2020\|EPI_ISL_751463\|2020-11 | ···················· | ···················· | ····················· | ···················· | ···················· | ···················· |
| AA | 2968 | hCoV-19/France/OCC-144/2020\|EPI_ISL_751464\|2020-11 | ···················· | ···················· | ····················· | ···················· | ···················· | ···················· |
| AA | 2969 | hCoV-19/France/OCC-145/2020\|EPI_ISL_751465\|2020-11 | ···················· | ···················· | ····················· | ···················· | ···················· | ···················· |
| AA | 2970 | hCoV-19/France/OCC-146/2020\|EPI_ISL_751466\|2020-11 | T··················· | ···················· | ····················· | ···················· | ···················· | ···················· |
| AA | 2971 | hCoV-19/France/OCC-159/2020\|EPI_ISL_751478\|2020-10 | ···················· | ···················· | ····················· | ···················· | ···················· | ···················· |
| AA | 2972 | hCoV-19/France/OCC-160/2020\|EPI_ISL_751479\|2020-10 | ···················· | ···················· | ····················· | ···················· | ···················· | ···················· |
| AA | 2973 | hCoV-19/France/OCC-161/2020\|EPI_ISL_751480\|2020-10 | ···················· | ···················· | ····················· | ···················· | ···················· | ···················· |
| AA | 2974 | hCoV-19/France/OCC-162/2020\|EPI_ISL_751481\|2020-10 | ···················· | ···················· | ····················· | ···················· | ···················· | ···················· |
| AA | 2975 | hCoV-19/France/OCC-163/2020\|EPI_ISL_751482\|2020-10 | ···················· | ···················· | ····················· | ···················· | ···················· | ···················· |
| AA | 2976 | hCoV-19/France/OCC-164/2020\|EPI_ISL_751483\|2020-10 | ···················· | ···················· | ····················· | ···················· | ···················· | ···················· |
| AA | 2977 | hCoV-19/France/OCC-165/2020\|EPI_ISL_751484\|2020-10 | ···················· | ···················· | ····················· | ···················· | ···················· | ···················· |
| AA | 2978 | hCoV-19/France/OCC-166/2020\|EPI_ISL_751485\|2020-10 | ···················· | ···················· | ····················· | ···················· | ···················· | ···················· |
| AA | 2979 | hCoV-19/France/OCC-167/2020\|EPI_ISL_751486\|2020-10 | ···················· | ···················· | ····················· | ···················· | ···················· | ···················· |
| AA | 2980 | hCoV-19/France/OCC-168/2020\|EPI_ISL_751487\|2020-10 | ···················· | ···················· | ····················· | ···················· | ···················· | ···················· |
| AA | 2981 | hCoV-19/France/OCC-169/2020\|EPI_ISL_751488\|2020-10 | ···················· | ···················· | ····················· | ···················· | ···················· | ···················· |
| AA | 2982 | hCoV-19/France/OCC-170/2020\|EPI_ISL_751489\|2020-10 | ···················· | ···················· | ····················· | ···················· | ···················· | ···················· |
| AA | 2983 | hCoV-19/France/OCC-171/2020\|EPI_ISL_751490\|2020-10 | ···················· | ···················· | ····················· | ···················· | ···················· | ···················· |
| AA | 2984 | hCoV-19/France/OCC-172/2020\|EPI_ISL_751491\|2020-11 | ···················· | ···················· | ····················· | ···················· | ···················· | ···················· |
| AA | 2985 | hCoV-19/France/OCC-173/2020\|EPI_ISL_751492\|2020-11 | ···················· | ···················· | ····················· | ···················· | ···················· | ···················· |
| AA | 2986 | hCoV-19/France/OCC-174/2020\|EPI_ISL_751493\|2020-11 | ···················· | ···················· | ····················· | ···················· | ···················· | ···················· |
| AA | 2987 | hCoV-19/France/OCC-175/2020\|EPI_ISL_751494\|2020-11 | ···················· | ···················· | ····················· | ···················· | ···················· | ···················· |
| AA | 2988 | hCoV-19/France/OCC-177/2020\|EPI_ISL_751495\|2020-10 | ···················· | ···················· | ····················· | ···················· | ···················· | ···················· |
| AA | 2989 | hCoV-19/France/OCC-178/2020\|EPI_ISL_751496\|2020-10 | ···················· | ···················· | ····················· | ···················· | ···················· | ···················· |
| AA | 2990 | hCoV-19/France/OCC-179/2020\|EPI_ISL_751497\|2020-11 | ···················· | ···················· | ····················· | ···················· | ···················· | ···················· |
| AA | 2991 | hCoV-19/France/OCC-180/2020\|EPI_ISL_751498\|2020-11 | ···················· | ···················· | ····················· | ···················· | ···················· | ···················· |
| AA | 2992 | hCoV-19/France/OCC-181/2020\|EPI_ISL_751499\|2020-10 | ···················· | ···················· | ····················· | ···················· | ···················· | ···················· |
| AA | 2993 | hCoV-19/USA/HI-H200137/2020\|EPI_ISL_752700\|2020-03 | ···················· | ·A·················· | ····················· | ···················· | ···················· | ···················· |
| AA | 2994 | hCoV-19/USA/HI-H200138/2020\|EPI_ISL_752701\|2020-07 | ···················· | ···················· | ····················· | ···················· | ···················· | ···················· |
| AA | 2995 | hCoV-19/USA/HI-H200139/2020\|EPI_ISL_752702\|2020-07 | ···················· | ···················· | ····················· | ···················· | ···················· | ···················· |
| AA | 2996 | hCoV-19/USA/HI-H200140/2020\|EPI_ISL_752703\|2020-07 | ···················· | ···················· | ····················· | ···················· | ···················· | ···················· |
| AA | 2997 | hCoV-19/USA/HI-H200141/2020\|EPI_ISL_752704\|2020-06 | ···················· | ···················· | ····················· | ···················· | ···················· | ···················· |
| AA | 2998 | hCoV-19/USA/HI-H200142/2020\|EPI_ISL_752705\|2020-06 | ···················· | ···················· | ····················· | ···················· | ···················· | ···················· |
| AA | 2999 | hCoV-19/USA/HI-H200143/2020\|EPI_ISL_752706\|2020-06 | ···················· | ···················· | ····················· | ···················· | ···················· | ···················· |
| AA | 3000 | hCoV-19/USA/HI-H200144/2020\|EPI_ISL_752707\|2020-06 | ···················· | ···················· | ····················· | ···················· | ···················· | ···················· |
| AA | 3001 | hCoV-19/USA/HI-H200145/2020\|EPI_ISL_752708\|2020-07 | ···················· | ···················· | ····················· | ···················· | ···················· | ···················· |
| AA | 3002 | hCoV-19/USA/HI-H200146/2020\|EPI_ISL_752709\|2020-07 | ···················· | ···················· | ····················· | ···················· | ···················· | ···················· |
| AA | 3003 | hCoV-19/USA/HI-H200147/2020\|EPI_ISL_752710\|2020-03 | ···················· | ·A·················· | ····················· | ···················· | ···················· | ···················· |
| AA | 3004 | hCoV-19/USA/HI-H200148/2020\|EPI_ISL_752711\|2020-03 | ···················· | ·A·················· | ····················· | ···················· | ···················· | ···················· |
| AA | 3005 | hCoV-19/USA/HI-H200149/2020\|EPI_ISL_752712\|2020-07 | ···················· | ···················· | ····················· | ···················· | ···················· | ···················· |
| AA | 3006 | hCoV-19/USA/HI-H200150/2020\|EPI_ISL_752713\|2020-07 | ···················· | ···················· | ····················· | ···················· | ···················· | ···················· |
| AA | 3007 | hCoV-19/USA/HI-H200151/2020\|EPI_ISL_752714\|2020-07 | ···················· | ···················· | ····················· | ···················· | ···················· | ···················· |
| AA | 3008 | hCoV-19/USA/HI-H200152/2020\|EPI_ISL_752715\|2020-07 | ···················· | ···················· | ····················· | ···················· | ···················· | ···················· |
| AA | 3009 | hCoV-19/USA/HI-H200153/2020\|EPI_ISL_752716\|2020-07 | ···················· | ···················· | ····················· | ···················· | ···················· | ···················· |
| AA | 3010 | hCoV-19/USA/HI-H200154/2020\|EPI_ISL_752717\|2020-07 | ···················· | ·A·················· | ····················· | ···················· | ···················· | ···················· |
| AA | 3011 | hCoV-19/USA/HI-H200155/2020\|EPI_ISL_752718\|2020-07 | ···················· | ···················· | ····················· | ···················· | ···················· | ···················· |
| AA | 3012 | hCoV-19/USA/HI-H200156/2020\|EPI_ISL_752719\|2020-07 | ···················· | ·A·················· | ····················· | ···················· | ···················· | ···················· |
| AA | 3013 | hCoV-19/USA/HI-H200158/2020\|EPI_ISL_752720\|2020-07 | ···················· | ···················· | ····················· | ···················· | ···················· | ···················· |
| AA | 3014 | hCoV-19/USA/HI-H200183/2020\|EPI_ISL_752743\|2020-07 | ···················· | ·A·················· | ····················· | ···················· | ···················· | ···················· |
| AA | 3015 | hCoV-19/USA/HI-H200184/2020\|EPI_ISL_752744\|2020-07 | ···················· | ·A·················· | ····················· | ···················· | ···················· | ···················· |
| AA | 3016 | hCoV-19/USA/HI-H200185/2020\|EPI_ISL_752745\|2020-07 | ···················· | ·A·················· | ····················· | ···················· | ···················· | ···················· |
| AA | 3017 | hCoV-19/USA/HI-H200187/2020\|EPI_ISL_752746\|2020-07 | ···················· | ·A·················· | ····················· | ···················· | ···················· | ···················· |
| AA | 3018 | hCoV-19/USA/HI-H200188/2020\|EPI_ISL_752747\|2020-08 | ···················· | ·A·················· | ····················· | ···················· | ···················· | ···················· |
| AA | 3019 | hCoV-19/USA/HI-H200189/2020\|EPI_ISL_752748\|2020-08 | ···················· | ·A·················· | ····················· | ···················· | ···················· | ···················· |
| AA | 3020 | hCoV-19/USA/HI-H200191/2020\|EPI_ISL_752750\|2020-08 | ···················· | ·A·················· | ····················· | ···················· | ···················· | ···················· |
| AA | 3021 | hCoV-19/USA/HI-H200192/2020\|EPI_ISL_752751\|2020-08 | ···················· | ·A·················· | ····················· | ···················· | ···················· | ···················· |
| AA | 3022 | hCoV-19/USA/HI-H200193/2020\|EPI_ISL_752752\|2020-08 | ···················· | ·A·················· | ····················· | ···················· | ···················· | ···················· |
| AA | 3023 | hCoV-19/USA/HI-H200194/2020\|EPI_ISL_752753\|2020-08 | ···················· | ·A·················· | ····················· | ···················· | ···················· | ···················· |
| AA | 3024 | hCoV-19/USA/HI-H200195/2020\|EPI_ISL_752754\|2020-08 | ···················· | ·A·················· | ····················· | ···················· | ···················· | ···················· |
| AA | 3025 | hCoV-19/USA/HI-H200196/2020\|EPI_ISL_752755\|2020-08 | ···················· | ·A·················· | ····················· | ···················· | ···················· | ···················· |
| AA | 3026 | hCoV-19/USA/HI-H200197/2020\|EPI_ISL_752756\|2020-08 | ···················· | ·A·················· | ····················· | ···················· | ···················· | ···················· |
| AA | 3027 | hCoV-19/USA/HI-H200198/2020\|EPI_ISL_752757\|2020-08 | ···················· | ·A·················· | ····················· | ···················· | ···················· | ···················· |
| AA | 3028 | hCoV-19/USA/HI-H200199/2020\|EPI_ISL_752758\|2020-08 | ···················· | ·A·················· | ····················· | ···················· | ···················· | ···················· |
| AA | 3029 | hCoV-19/USA/HI-H200200/2020\|EPI_ISL_752759\|2020-08 | ···················· | ·A·················· | ····················· | ···················· | ···················· | ···················· |
| AA | 3030 | hCoV-19/USA/HI-H200201/2020\|EPI_ISL_752760\|2020-08 | ···················· | ·A·················· | ····················· | ···················· | ···················· | ···················· |
| AA | 3031 | hCoV-19/USA/HI-H200202/2020\|EPI_ISL_752761\|2020-08 | ···················· | ·A·················· | ····················· | ···················· | ···················· | ···················· |
| AA | 3032 | hCoV-19/USA/HI-H200203/2020\|EPI_ISL_752762\|2020-08 | ···················· | ·A·················· | ····················· | ···················· | ···················· | ···················· |
| AA | 3033 | hCoV-19/USA/HI-H200204/2020\|EPI_ISL_752763\|2020-08 | ···················· | ·A·················· | ····················· | ···················· | ···················· | ···················· |
| AA | 3034 | hCoV-19/USA/HI-H200205/2020\|EPI_ISL_752764\|2020-08 | ···················· | ·A·················· | ····················· | ···················· | ···················· | ···················· |
| AA | 3035 | hCoV-19/USA/HI-H200206/2020\|EPI_ISL_752765\|2020-08 | ···················· | ·A·················· | ····················· | ···················· | ···················· | ···················· |
| AA | 3036 | hCoV-19/USA/HI-H200207/2020\|EPI_ISL_752766\|2020-08 | ···················· | ·A·················· | ····················· | ···················· | ···················· | ···················· |
| AA | 3037 | hCoV-19/USA/HI-H200208/2020\|EPI_ISL_752767\|2020-08 | ···················· | ·A·················· | ····················· | ···················· | ···················· | ···················· |
| AA | 3038 | hCoV-19/USA/HI-H200209/2020\|EPI_ISL_752768\|2020-08 | ···················· | ·A·················· | ····················· | ···················· | ···················· | ···················· |
| AA | 3039 | hCoV-19/USA/HI-H200210/2020\|EPI_ISL_752769\|2020-08 | ···················· | ·A·················· | ····················· | ···················· | ···················· | ···················· |
| AA | 3040 | hCoV-19/USA/HI-H200211/2020\|EPI_ISL_752770\|2020-08 | ···················· | ·A·················· | ····················· | ···················· | ···················· | ···················· |
| AA | 3041 | hCoV-19/USA/HI-H200212/2020\|EPI_ISL_752771\|2020-08 | ···················· | ·A·················· | ····················· | ···················· | ···················· | ···················· |
| AA | 3042 | hCoV-19/USA/HI-H200213/2020\|EPI_ISL_752772\|2020-08 | ···················· | ·A·················· | ····················· | ···················· | ···················· | ···················· |
| AA | 3043 | hCoV-19/USA/HI-H200214/2020\|EPI_ISL_752773\|2020-08 | ···················· | ·A·················· | ····················· | ···················· | ···················· | ···················· |

AA 3044 hCoV-19/USA/HI-H200215/2020|EPI_ISL_752774|2020-08 ···················· ·A·················· ···················· ···················· ···················· ····················
AA 3045 hCoV-19/USA/HI-H200216/2020|EPI_ISL_752775|2020-08 ···················· ·A·················· ···················· ···················· ···················· ····················
AA 3046 hCoV-19/USA/HI-H200217/2020|EPI_ISL_752776|2020-08 ···················· ·A·················· ···················· ···················· ···················· ····················
AA 3047 hCoV-19/USA/HI-H200218/2020|EPI_ISL_752777|2020-08 ···················· ·A·················· ···················· ···················· ···················· ····················
AA 3048 hCoV-19/USA/HI-H200219/2020|EPI_ISL_752778|2020-08 ···················· ·A·················· ···················· ···················· ···················· ····················
AA 3049 hCoV-19/USA/HI-H200220/2020|EPI_ISL_752779|2020-08 ···················· ·A·················· ···················· ···················· ···················· ····················
AA 3050 hCoV-19/USA/HI-H200221/2020|EPI_ISL_752780|2020-08 ···················· ·A·················· ···················· ···················· ···················· ····················
AA 3051 hCoV-19/USA/HI-H200222/2020|EPI_ISL_752781|2020-08 ···················· ·A·················· ···················· ···················· ···················· ····················
AA 3052 hCoV-19/USA/HI-H200223/2020|EPI_ISL_752782|2020-08 ···················· ·A·················· ···················· ···················· ···················· ····················
AA 3053 hCoV-19/USA/HI-H200224/2020|EPI_ISL_752783|2020-08 ···················· ·A·················· ···················· ···················· ···················· ····················
AA 3054 hCoV-19/USA/HI-H200225/2020|EPI_ISL_752784|2020-08 ···················· ·A·················· ···················· ···················· ···················· ····················
AA 3055 hCoV-19/USA/HI-H200226/2020|EPI_ISL_752785|2020-08 ···················· ·A·················· ···················· ···················· ···················· ····················
AA 3056 hCoV-19/USA/HI-H200227/2020|EPI_ISL_752786|2020-08 ···················· ·A·················· ···················· ···················· ···················· ····················
AA 3057 hCoV-19/USA/HI-H200228/2020|EPI_ISL_752787|2020-08 ···················· ·A·················· ···················· ···················· ···················· ····················
AA 3058 hCoV-19/USA/HI-H200229/2020|EPI_ISL_752788|2020-08 ···················· ·A·················· ···················· ···················· ···················· ····················
AA 3059 hCoV-19/USA/HI-H200230/2020|EPI_ISL_752789|2020-08 ···················· ·A·················· ···················· ···················· ···················· ····················
AA 3060 hCoV-19/USA/HI-H200234/2020|EPI_ISL_752793|2020-08 ···················· ·A·················· ···················· ···················· ···················· ····················
AA 3061 hCoV-19/USA/HI-H200235/2020|EPI_ISL_752794|2020-08 ···················· ·A·················· ···················· ···················· ···················· ····················
AA 3062 hCoV-19/USA/HI-H200236/2020|EPI_ISL_752795|2020-08 ···················· ·A·················· ···················· ···················· ···················· ····················
AA 3063 hCoV-19/USA/HI-H200237/2020|EPI_ISL_752796|2020-08 ···················· ·A·················· ···················· ···················· ···················· ····················
AA 3064 hCoV-19/USA/HI-H200238/2020|EPI_ISL_752797|2020-08 ···················· ·A·················· ···················· ···················· ···················· ····················
AA 3065 hCoV-19/Finland/18SS20D3/2020|EPI_ISL_756290|2020- ···················· ·A·················· ···················· ···················· ···················· ··A·················
AA 3066 hCoV-19/Finland/18SS6E7/2020|EPI_ISL_756291|2020-0 ···················· ·A·················· ···················· ···················· ···················· ··A·················
AA 3067 hCoV-19/Finland/18SS6F5/2020|EPI_ISL_756292|2020-0 ···················· ·A·················· ···················· ···················· ···················· ··A·················
AA 3068 hCoV-19/Brazil/BA-1/2020|EPI_ISL_756293|2020-06-01 ···················· ···················· ···················· ···················· ···················· ····················
AA 3069 hCoV-19/Brazil/BA-2/2020|EPI_ISL_756294|2020-10-26 ···················· ···················· ···················· ···················· ···················· ····················
AA 3070 hCoV-19/Denmark/DCGC-20519/2020|EPI_ISL_758900|202 ···················· ···················· ···················· ···················· ···················· ····················
AA 3071 hCoV-19/Denmark/DCGC-20520/2020|EPI_ISL_758901|202 ···················· ···················· ···················· ···················· ···················· ····················
AA 3072 hCoV-19/Denmark/DCGC-20524/2020|EPI_ISL_758902|202 ···················· ···················· ···················· ···················· ···················· ····················
AA 3073 hCoV-19/Denmark/DCGC-18698/2020|EPI_ISL_750131|202 ···················· ···················· ···················· ···················· ·····T·············· ····················
AA 3074 hCoV-19/Denmark/DCGC-18710/2020|EPI_ISL_750136|202 ···················· ···················· ···················· ···················· ···················· ····················
AA 3075 hCoV-19/Denmark/DCGC-18711/2020|EPI_ISL_750137|202 ···················· ···················· ···················· ···················· ···················· ····················
AA 3076 hCoV-19/Denmark/DCGC-18713/2020|EPI_ISL_750138|202 ···················· ···················· ···················· ···················· ···················· ····················
AA 3077 hCoV-19/Denmark/DCGC-18714/2020|EPI_ISL_750139|202 ···················· ···················· ···················· ···················· ···················· ····················
AA 3078 hCoV-19/Denmark/DCGC-18716/2020|EPI_ISL_750140|202 ···················· ···················· ···················· ···················· ···················· ····················
AA 3079 hCoV-19/Denmark/DCGC-18718/2020|EPI_ISL_750141|202 ···················· ···················· ···················· ···················· ···················· ····················
AA 3080 hCoV-19/Denmark/DCGC-18719/2020|EPI_ISL_750142|202 ···················· ···················· ···················· ···················· ···················· ····················
AA 3081 hCoV-19/Denmark/DCGC-18720/2020|EPI_ISL_750143|202 ···················· ···················· ···················· ···················· ···················· ····················
AA 3082 hCoV-19/Denmark/DCGC-18722/2020|EPI_ISL_750144|202 ···················· ···················· ···················· ···················· ···················· ····················
AA 3083 hCoV-19/Denmark/DCGC-18724/2020|EPI_ISL_750145|202 ···················· ···················· ···················· ···················· ···················· ····················
AA 3084 hCoV-19/Denmark/DCGC-18725/2020|EPI_ISL_750146|202 ···················· ···················· ···················· ···················· ···················· ····················
AA 3085 hCoV-19/France/OCC-140/2020|EPI_ISL_751460|2020-11 ···················· ···················· ···················· ···················· ···················· ····················
AA 3086 hCoV-19/France/OCC-142/2020|EPI_ISL_751462|2020-11 ···················· ···················· ···················· ···················· ···················· ····················
AA 3087 hCoV-19/France/OCC-147/2020|EPI_ISL_751467|2020-11 ···················· ···················· ···················· ···················· ···················· ····················
AA 3088 hCoV-19/France/OCC-148/2020|EPI_ISL_751468|2020-11 ···················· ···················· ···················· ···················· ···················· ····················
AA 3089 hCoV-19/France/OCC-149/2020|EPI_ISL_751469|2020-11 ···················· ···················· ···················· ···················· ···················· ····················
AA 3090 hCoV-19/France/OCC-150/2020|EPI_ISL_751470|2020-11 ···················· ···················· ···················· ···················· ···················· ····················
AA 3091 hCoV-19/France/OCC-151/2020|EPI_ISL_751471|2020-11 ···················· ···················· ···················· ···················· ···················· ····················
AA 3092 hCoV-19/France/OCC-152/2020|EPI_ISL_751472|2020-11 ···················· ···················· ···················· ···················· ···················· ····················
AA 3093 hCoV-19/France/OCC-153/2020|EPI_ISL_751473|2020-11 ···················· ···················· ···················· ···················· ···················· ····················
AA 3094 hCoV-19/France/OCC-154/2020|EPI_ISL_751474|2020-11 ···················· ···················· ···················· ···················· ···················· ····················
AA 3095 hCoV-19/France/OCC-155/2020|EPI_ISL_751475|2020-11 ···················· ···················· ···················· ···················· ···················· ····················
AA 3096 hCoV-19/France/OCC-156/2020|EPI_ISL_751476|2020-11 ···················· ···················· ···················· ···················· ···················· ····················
AA 3097 hCoV-19/France/OCC-158/2020|EPI_ISL_751477|2020-10 ···················· ···················· ···················· ···················· ···················· ····················
AA 3098 hCoV-19/USA/HI-H200231/2020|EPI_ISL_752790|2020-08 ···················· ·A·················· ···················· ···················· ···················· ····················
AA 3099 hCoV-19/USA/HI-H200232/2020|EPI_ISL_752791|2020-08 ···················· ·A·················· ···················· ···················· ···················· ····················
AA 3100 hCoV-19/USA/HI-H200233/2020|EPI_ISL_752792|2020-08 ···················· ·A·················· ···················· ···················· ···················· ····················
AA 3101 hCoV-19/USA/HI-H200239/2020|EPI_ISL_752798|2020-08 ···················· ·A·················· ···················· ···················· ···················· ····················
AA 3102 hCoV-19/USA/HI-H200240/2020|EPI_ISL_752799|2020-08 ···················· ·A·················· ···················· ···················· ···················· ····················
AA 3103 hCoV-19/USA/CA-CDPH294/2020|EPI_ISL_755015|2020-12 ···················· ···················· ···················· ···················· ···················· ····················
AA 3104 hCoV-19/USA/CA-CDPH068/2020|EPI_ISL_755040|2020-04 ···················· ···················· ···················· ···················· ···················· ····················
AA 3105 hCoV-19/USA/CA-CDPH069/2020|EPI_ISL_755041|2020-04 ···················· ···················· ···················· ···················· ···················· ····················
AA 3106 hCoV-19/USA/CA-CDPH072/2020|EPI_ISL_755042|2020-07 ···················· ···················· ···················· ···················· ···················· ····················
AA 3107 hCoV-19/USA/CA-CDPH079/2020|EPI_ISL_755043|2020-07 ···················· ···················· ···················· ···················· ···················· ····················
AA 3108 hCoV-19/USA/CA-CDPH084/2020|EPI_ISL_755044|2020-07 ···················· ···················· ···················· ···················· ···················· ····················
AA 3109 hCoV-19/USA/CA-CDPH085/2020|EPI_ISL_755045|2020-07 ···················· ···················· ···················· ···················· ···················· ····················
AA 3110 hCoV-19/USA/CA-CDPH086/2020|EPI_ISL_755046|2020-07 ···················· ···················· ···················· ···················· ···················· ····················
AA 3111 hCoV-19/USA/CA-CDPH088/2020|EPI_ISL_755047|2020-08 ···················· ·A·················· ···················· ···················· ···················· ····················
AA 3112 hCoV-19/USA/CA-CDPH093/2020|EPI_ISL_755048|2020-08 ···················· ·A·················· ···················· ···················· ···················· ····················
AA 3113 hCoV-19/USA/CA-CDPH107/2020|EPI_ISL_755049|2020-09 ···················· ·A·················· ···················· ···················· ···················· ····················
AA 3114 hCoV-19/USA/CA-CDPH110/2020|EPI_ISL_755050|2020-09 ···················· ·A·················· ···················· ···················· ···················· ····················
AA 3115 hCoV-19/USA/CA-CDPH111/2020|EPI_ISL_755051|2020-09 ···················· ·A·················· ···················· ···················· ···················· ····················
AA 3116 hCoV-19/USA/CA-CDPH112/2020|EPI_ISL_755052|2020-09 ···················· ···················· ···················· ···················· ···················· ····················
AA 3117 hCoV-19/USA/CA-CDPH113/2020|EPI_ISL_755053|2020-09 ···················· ···················· ···················· ···················· ···················· ····················
AA 3118 hCoV-19/USA/CA-CDPH130/2020|EPI_ISL_755054|2020-07 ···················· ···················· ···················· ···················· ···················· ····················
AA 3119 hCoV-19/USA/CA-CDPH131/2020|EPI_ISL_755055|2020-07 ···················· ···················· ···················· ···················· ···················· ····················
AA 3120 hCoV-19/USA/CA-CDPH134/2020|EPI_ISL_755056|2020-07 ···················· ···················· ···················· ···················· ···················· ····················
AA 3121 hCoV-19/USA/CA-CDPH142/2020|EPI_ISL_755057|2020-09 ···················· ·A·················· ···················· ···················· ···················· ····················
AA 3122 hCoV-19/USA/CA-CDPH151/2020|EPI_ISL_755058|2020-09 ···················· ·A·················· ···················· ···················· ···················· ····················
AA 3123 hCoV-19/USA/CA-CDPH152/2020|EPI_ISL_755059|2020-09 ···················· ·A·················· ···················· ···················· ···················· ····················

| | | | | | | | |
|---|---|---|---|---|---|---|---|
| AA | 3124 hCoV-19/USA/CA-CDPH160/2020\|EPI_ISL_755060\|2020-06 | ···················· | ···················· | ····················· | ···················· | ···················· | ···················· |
| AA | 3125 hCoV-19/USA/CA-CDPH167/2020\|EPI_ISL_755061\|2020-06 | ···················· | ···················· | ····················· | ···················· | ···················· | ···················· |
| AA | 3126 hCoV-19/USA/CA-CDPH206/2020\|EPI_ISL_755062\|2020-11 | ···················· | ···················· | ····················· | ···················· | ···················· | ···················· |
| AA | 3127 hCoV-19/USA/CA-CDPH207/2020\|EPI_ISL_755063\|2020-11 | ···················· | ···················· | ····················· | ···················· | ···················· | ···················· |
| AA | 3128 hCoV-19/USA/CA-CDPH208/2020\|EPI_ISL_755064\|2020-11 | ···················· | ···················· | ····················· | ···················· | ···················· | ···················· |
| AA | 3129 hCoV-19/USA/CA-CDPH209/2020\|EPI_ISL_755065\|2020-10 | ···················· | ···················· | ····················· | ···················· | ···················· | ···················· |
| AA | 3130 hCoV-19/USA/CA-CDPH210/2020\|EPI_ISL_755066\|2020-10 | ···················· | ···················· | ····················· | ···················· | ···················· | ···················· |
| AA | 3131 hCoV-19/USA/CA-CDPH211/2020\|EPI_ISL_755067\|2020-11 | ···················· | ···················· | ····················· | ···················· | ···················· | ···················· |
| AA | 3132 hCoV-19/USA/CA-CDPH212/2020\|EPI_ISL_755068\|2020-11 | ···················· | ···················· | ····················· | ···················· | ···················· | ···················· |
| AA | 3133 hCoV-19/USA/CA-CDPH213/2020\|EPI_ISL_755069\|2020-11 | ···················· | ···················· | ····················· | ···················· | ···················· | ···················· |
| AA | 3134 hCoV-19/USA/CA-CDPH214/2020\|EPI_ISL_755070\|2020-11 | ···················· | ···················· | ····················· | ···················· | ···················· | ···················· |
| AA | 3135 hCoV-19/USA/CA-CDPH215/2020\|EPI_ISL_755071\|2020-11 | ···················· | ···················· | ····················· | ···················· | ···················· | ···················· |
| AA | 3136 hCoV-19/USA/CA-CDPH216/2020\|EPI_ISL_755072\|2020-11 | ···················· | ···················· | ····················· | ···················· | ···················· | ···················· |
| AA | 3137 hCoV-19/USA/CA-CDPH217/2020\|EPI_ISL_755073\|2020-11 | ···················· | ···················· | ····················· | ···················· | ···················· | ···················· |
| AA | 3138 hCoV-19/USA/CA-CDPH222/2020\|EPI_ISL_755074\|2020-11 | ···················· | ···················· | ····················· | ···················· | ···················· | ···················· |
| AA | 3139 hCoV-19/USA/CA-CDPH223/2020\|EPI_ISL_755075\|2020-11 | ···················· | ···················· | ····················· | ···················· | ···················· | ···················· |
| AA | 3140 hCoV-19/USA/CA-CDPH225/2020\|EPI_ISL_755076\|2020-11 | ···················· | ···················· | ····················· | ···················· | ···················· | ···················· |
| AA | 3141 hCoV-19/USA/CA-CDPH226/2020\|EPI_ISL_755077\|2020-11 | ···················· | ···················· | ····················· | ···················· | ···················· | ···················· |
| AA | 3142 hCoV-19/USA/CA-CDPH227/2020\|EPI_ISL_755078\|2020-11 | ···················· | ···················· | ····················· | ···················· | ···················· | ···················· |
| AA | 3143 hCoV-19/USA/CA-CDPH229/2020\|EPI_ISL_755079\|2020-11 | ···················· | ···················· | ····················· | ···················· | ···················· | ···················· |
| AA | 3144 hCoV-19/USA/WA-UW-46859/2020\|EPI_ISL_756302\|2020-1 | ···················· | ···················· | ····················· | ···················· | ···················· | ···················· |
| AA | 3145 hCoV-19/USA/WA-UW-46995/2020\|EPI_ISL_756303\|2020-1 | ···················· | ·A·················· | ····················· | ···················· | ···················· | ···················· |
| AA | 3146 hCoV-19/USA/WA-UW-47072/2020\|EPI_ISL_756304\|2020-1 | ···················· | ···················· | ····················· | ···················· | ···················· | ···················· |
| AA | 3147 hCoV-19/USA/WA-UW-47108/2020\|EPI_ISL_756305\|2020-1 | ···················· | ···················· | ····················· | ···················· | ···················· | ···················· |
| AA | 3148 hCoV-19/USA/CA-IGI-0384/2020\|EPI_ISL_756312\|2020-0 | ···················· | ·A·················· | ····················· | ···················· | ···················· | ···················· |
| AA | 3149 hCoV-19/USA/CA-IGI-0390/2020\|EPI_ISL_756313\|2020-0 | ···················· | ···················· | ····················· | ···················· | ···················· | ···················· |
| AA | 3150 hCoV-19/USA/CA-IGI-0393/2020\|EPI_ISL_756314\|2020-0 | ···················· | ·A·················· | ····················· | ···················· | ···················· | ···················· |
| AA | 3151 hCoV-19/USA/CA-IGI-0395/2020\|EPI_ISL_756315\|2020-0 | ···················· | ···················· | ····················· | ···················· | ···················· | ···················· |
| AA | 3152 hCoV-19/USA/CA-IGI-0398/2020\|EPI_ISL_756316\|2020-0 | ···················· | ···················· | ····················· | ···················· | ···················· | ···················· |
| AA | 3153 hCoV-19/USA/CA-IGI-0403/2020\|EPI_ISL_756317\|2020-0 | ···················· | ···················· | ····················· | ···················· | ···················· | ···················· |
| AA | 3154 hCoV-19/USA/CA-IGI-0406/2020\|EPI_ISL_756318\|2020-0 | ···················· | ·A·················· | ····················· | ···················· | ···················· | ···················· |
| AA | 3155 hCoV-19/USA/CA-IGI-0414/2020\|EPI_ISL_756320\|2020-0 | ···················· | ···················· | ····················· | ···················· | ···················· | ···················· |
| AA | 3156 hCoV-19/USA/CA-IGI-0467/2020\|EPI_ISL_756330\|2020-0 | ···················· | ···················· | ····················· | ···················· | ···················· | ···················· |
| AA | 3157 hCoV-19/USA/CA-IGI-0509/2020\|EPI_ISL_756332\|2020-0 | ···················· | ···················· | ····················· | ···················· | ···················· | ···················· |
| AA | 3158 hCoV-19/USA/CA-IGI-0704/2020\|EPI_ISL_756336\|2020-0 | ···················· | ···················· | ····················· | ···················· | ···················· | ···················· |
| AA | 3159 hCoV-19/USA/CA-IGI-0798/2020\|EPI_ISL_756339\|2020-0 | ···················· | ···················· | ····················· | ···················· | ···················· | ···················· |
| AA | 3160 hCoV-19/USA/CA-IGI-0803/2020\|EPI_ISL_756340\|2020-0 | ···················· | ···················· | ····················· | ···················· | ···················· | ···················· |
| AA | 3161 hCoV-19/USA/CA-IGI-0806/2020\|EPI_ISL_756342\|2020-0 | ···················· | ···················· | ····················· | ···················· | ···················· | ···················· |
| AA | 3162 hCoV-19/USA/CA-IGI-0807/2020\|EPI_ISL_756343\|2020-0 | ···················· | ···················· | ····················· | ···················· | ···················· | ···················· |
| AA | 3163 hCoV-19/USA/CA-IGI-0814/2020\|EPI_ISL_756346\|2020-0 | ···················· | ···················· | ····················· | ···················· | ···················· | ···················· |
| AA | 3164 hCoV-19/USA/CA-IGI-0819/2020\|EPI_ISL_756347\|2020-0 | ···················· | ···················· | ····················· | ···················· | ···················· | ···················· |
| AA | 3165 hCoV-19/USA/CA-IGI-0821/2020\|EPI_ISL_756348\|2020-0 | ···················· | ···················· | ····················· | ···················· | ···················· | ···················· |
| AA | 3166 hCoV-19/USA/CA-IGI-0824/2020\|EPI_ISL_756349\|2020-0 | ···················· | ···················· | ····················· | ···················· | ···················· | ···················· |
| AA | 3167 hCoV-19/Thailand/Bangkok-Si11/2020\|EPI_ISL_756375\| | ···················· | ·A·················· | ····················· | ···················· | ···················· | ··A················· |
| AA | 3168 hCoV-19/Indonesia/JB-NIHRDC-0056536/2020\|EPI_ISL_7 | ···················· | ···················· | ····················· | ···················· | ···················· | ···················· |
| AA | 3169 hCoV-19/England/MILK-A8E255/2020\|EPI_ISL_756377\|20 | ···················· | ···················· | ····················· | ···················· | ···················· | ···················· |
| AA | 3170 hCoV-19/England/ALDP-C8962F/2020\|EPI_ISL_756378\|20 | ···················· | ···················· | ····················· | ···················· | ···················· | ···················· |
| AA | 3171 hCoV-19/England/ALDP-C8B5C6/2020\|EPI_ISL_756382\|20 | ···················· | ···················· | ····················· | ···················· | ···················· | ···················· |
| AA | 3172 hCoV-19/England/ALDP-C88EEC/2020\|EPI_ISL_756385\|20 | ···················· | ···················· | ····················· | ···················· | ···················· | ···················· |
| AA | 3173 hCoV-19/England/CAMC-C84EC2/2020\|EPI_ISL_756389\|20 | ···················· | ···················· | ····················· | ···················· | ·····T·············· | ···················· |
| AA | 3174 hCoV-19/England/ALDP-C88D1C/2020\|EPI_ISL_756390\|20 | ···················· | ···················· | ····················· | ···················· | ···················· | ···················· |
| AA | 3175 hCoV-19/England/CAMC-C85FBF/2020\|EPI_ISL_756392\|20 | ···················· | ···················· | ····················· | ···················· | ···················· | ···················· |
| AA | 3176 hCoV-19/England/CAMC-C86A17/2020\|EPI_ISL_756395\|20 | ···················· | ···················· | ····················· | ···················· | ···················· | ···················· |
| AA | 3177 hCoV-19/England/ALDP-C8B405/2020\|EPI_ISL_756398\|20 | ···················· | ···················· | ····················· | ···················· | ···················· | ···················· |
| AA | 3178 hCoV-19/Denmark/DCGC-22506/2020\|EPI_ISL_757600\|202 | ···················· | ···················· | ····················· | ···················· | ···················· | ···················· |
| AA | 3179 hCoV-19/Denmark/DCGC-22507/2020\|EPI_ISL_757601\|202 | ···················· | ···················· | ····················· | ···················· | ···················· | ···················· |
| AA | 3180 hCoV-19/Denmark/DCGC-22508/2020\|EPI_ISL_757602\|202 | ···················· | ···················· | ····················· | ···················· | ···················· | ···················· |
| AA | 3181 hCoV-19/Denmark/DCGC-22509/2020\|EPI_ISL_757603\|202 | ···················· | ···················· | ····················· | ···················· | ···················· | ···················· |
| AA | 3182 hCoV-19/Denmark/DCGC-22510/2020\|EPI_ISL_757604\|202 | ···················· | ···················· | ····················· | ···················· | ···················· | ···················· |
| AA | 3183 hCoV-19/Denmark/DCGC-22511/2020\|EPI_ISL_757605\|202 | ···················· | ···················· | ····················· | ···················· | ···················· | ···················· |
| AA | 3184 hCoV-19/Denmark/DCGC-22514/2020\|EPI_ISL_757606\|202 | ···················· | ···················· | ····················· | ···················· | ···················· | ···················· |
| AA | 3185 hCoV-19/Denmark/DCGC-22515/2020\|EPI_ISL_757607\|202 | ···················· | ···················· | ····················· | ···················· | ···················· | ···················· |
| AA | 3186 hCoV-19/Denmark/DCGC-22517/2020\|EPI_ISL_757608\|202 | ···················· | ···················· | ····················· | ···················· | ···················· | ···················· |
| AA | 3187 hCoV-19/Denmark/DCGC-22518/2020\|EPI_ISL_757609\|202 | ···················· | ···················· | ····················· | ···················· | ···················· | ···················· |
| AA | 3188 hCoV-19/Denmark/DCGC-22519/2020\|EPI_ISL_757610\|202 | ···················· | ···················· | ····················· | ···················· | ···················· | ···················· |
| AA | 3189 hCoV-19/Denmark/DCGC-22520/2020\|EPI_ISL_757611\|202 | ···················· | ···················· | ····················· | ···················· | ···················· | ···················· |
| AA | 3190 hCoV-19/Denmark/DCGC-22521/2020\|EPI_ISL_757612\|202 | ···················· | ···················· | ····················· | ···················· | ···················· | ···················· |
| AA | 3191 hCoV-19/Denmark/DCGC-22522/2020\|EPI_ISL_757613\|202 | ···················· | ···················· | ····················· | ···················· | ···················· | ···················· |
| AA | 3192 hCoV-19/Denmark/DCGC-22526/2020\|EPI_ISL_757614\|202 | ···················· | ···················· | ····················· | ···················· | ···················· | ···················· |
| AA | 3193 hCoV-19/Denmark/DCGC-22527/2020\|EPI_ISL_757615\|202 | ···················· | ···················· | ····················· | ···················· | ···················· | ···················· |
| AA | 3194 hCoV-19/Denmark/DCGC-22528/2020\|EPI_ISL_757616\|202 | ···················· | ···················· | ····················· | ···················· | ···················· | ···················· |
| AA | 3195 hCoV-19/Denmark/DCGC-22529/2020\|EPI_ISL_757617\|202 | ···················· | ···················· | ····················· | ···················· | ···················· | ···················· |
| AA | 3196 hCoV-19/Denmark/DCGC-22530/2020\|EPI_ISL_757618\|202 | ···················· | ···················· | ····················· | ···················· | ···················· | ···················· |
| AA | 3197 hCoV-19/Denmark/DCGC-22531/2020\|EPI_ISL_757619\|202 | ···················· | ···················· | ····················· | ···················· | ···················· | ···················· |
| AA | 3198 hCoV-19/Denmark/DCGC-22532/2020\|EPI_ISL_757620\|202 | ···················· | ···················· | ····················· | ···················· | ···················· | ···················· |
| AA | 3199 hCoV-19/Denmark/DCGC-22533/2020\|EPI_ISL_757621\|202 | ···················· | ···················· | ····················· | ···················· | ···················· | ···················· |
| AA | 3200 hCoV-19/Denmark/DCGC-22536/2020\|EPI_ISL_757622\|202 | ···················· | ···················· | ····················· | ···················· | ···················· | ···················· |
| AA | 3201 hCoV-19/Denmark/DCGC-22537/2020\|EPI_ISL_757623\|202 | ···················· | ·········A·········· | ····················· | ···················· | ···················· | ···················· |
| AA | 3202 hCoV-19/Denmark/DCGC-22538/2020\|EPI_ISL_757624\|202 | ···················· | ···················· | ····················· | ···················· | ···················· | ···················· |
| AA | 3203 hCoV-19/Denmark/DCGC-22539/2020\|EPI_ISL_757625\|202 | ···················· | ···················· | ····················· | ···················· | ···················· | ···················· |

| | | | | | | | | |
|---|---|---|---|---|---|---|---|---|
| AA | 3204 | hCoV-19/Denmark/DCGC-22540/2020\|EPI_ISL_757626\|202 | .................... | .................... | .................... | .................... | .................... | .................... |
| AA | 3205 | hCoV-19/Denmark/DCGC-22542/2020\|EPI_ISL_757627\|202 | .................... | .................... | .................... | .................... | .................... | .................... |
| AA | 3206 | hCoV-19/Denmark/DCGC-22543/2020\|EPI_ISL_757628\|202 | .................... | .................... | .................... | .................... | .................... | .................... |
| AA | 3207 | hCoV-19/Denmark/DCGC-22546/2020\|EPI_ISL_757629\|202 | .................... | .................... | .................... | .................... | .................... | .................... |
| AA | 3208 | hCoV-19/Denmark/DCGC-22548/2020\|EPI_ISL_757630\|202 | .................... | .................... | .................... | .................... | .................... | .................... |
| AA | 3209 | hCoV-19/Denmark/DCGC-22552/2020\|EPI_ISL_757631\|202 | .................... | .................... | .................... | .................... | .................... | .................... |
| AA | 3210 | hCoV-19/Denmark/DCGC-22553/2020\|EPI_ISL_757632\|202 | .................... | .................... | .................... | .................... | .................... | .................... |
| AA | 3211 | hCoV-19/Denmark/DCGC-22555/2020\|EPI_ISL_757633\|202 | .................... | .................... | .................... | .................... | .................... | .................... |
| AA | 3212 | hCoV-19/Denmark/DCGC-22557/2020\|EPI_ISL_757634\|202 | .................... | .................... | .................... | .................... | .................... | .................... |
| AA | 3213 | hCoV-19/Denmark/DCGC-22558/2020\|EPI_ISL_757635\|202 | .................... | .................... | .................... | .................... | .................... | .................... |
| AA | 3214 | hCoV-19/Denmark/DCGC-22560/2020\|EPI_ISL_757636\|202 | .................... | .................... | .................... | .................... | .................... | .................... |
| AA | 3215 | hCoV-19/Denmark/DCGC-22561/2020\|EPI_ISL_757637\|202 | .................... | .................... | .................... | .................... | .................... | .................... |
| AA | 3216 | hCoV-19/Denmark/DCGC-22563/2020\|EPI_ISL_757638\|202 | .................... | .................... | .................... | .................... | .................... | .................... |
| AA | 3217 | hCoV-19/Denmark/DCGC-22564/2020\|EPI_ISL_757639\|202 | .................... | .................... | .................... | .................... | .................... | .................... |
| AA | 3218 | hCoV-19/Denmark/DCGC-22565/2020\|EPI_ISL_757640\|202 | .................... | .................... | .................... | .................... | .................... | .................... |
| AA | 3219 | hCoV-19/Denmark/DCGC-22566/2020\|EPI_ISL_757641\|202 | .................... | .................... | .................... | .................... | .................... | .................... |
| AA | 3220 | hCoV-19/Denmark/DCGC-22567/2020\|EPI_ISL_757642\|202 | .................... | .................... | .................... | .................... | .................... | .................... |
| AA | 3221 | hCoV-19/Denmark/DCGC-22568/2020\|EPI_ISL_757643\|202 | .................... | .................... | .................... | .................... | .................... | .................... |
| AA | 3222 | hCoV-19/Denmark/DCGC-22570/2020\|EPI_ISL_757644\|202 | .................... | .................... | .................... | .................... | .................... | .................... |
| AA | 3223 | hCoV-19/Denmark/DCGC-22571/2020\|EPI_ISL_757645\|202 | .................... | .................... | .................... | .................... | .................... | .................... |
| AA | 3224 | hCoV-19/Denmark/DCGC-22572/2020\|EPI_ISL_757646\|202 | .................... | .................... | .................... | .................... | .................... | .................... |
| AA | 3225 | hCoV-19/Denmark/DCGC-22573/2020\|EPI_ISL_757647\|202 | .................... | .................... | .................... | .................... | .................... | .................... |
| AA | 3226 | hCoV-19/Denmark/DCGC-22574/2020\|EPI_ISL_757648\|202 | .................... | .................... | .................... | .................... | .................... | .................... |
| AA | 3227 | hCoV-19/Denmark/DCGC-22575/2020\|EPI_ISL_757649\|202 | .................... | .................... | .................... | .................... | .................... | .................... |
| AA | 3228 | hCoV-19/Denmark/DCGC-22576/2020\|EPI_ISL_757650\|202 | .................... | .................... | .................... | .................... | .................... | .................... |
| AA | 3229 | hCoV-19/Denmark/DCGC-22577/2020\|EPI_ISL_757651\|202 | .................... | .................... | .................... | .................... | .................... | .................... |
| AA | 3230 | hCoV-19/Denmark/DCGC-22579/2020\|EPI_ISL_757652\|202 | .................... | .................... | .................... | .................... | .................... | .................... |
| AA | 3231 | hCoV-19/Denmark/DCGC-22582/2020\|EPI_ISL_757653\|202 | .................... | .................... | .................... | .................... | .................... | .................... |
| AA | 3232 | hCoV-19/Denmark/DCGC-22586/2020\|EPI_ISL_757654\|202 | .................... | .................... | .................... | .................... | .................... | .................... |
| AA | 3233 | hCoV-19/Denmark/DCGC-22587/2020\|EPI_ISL_757655\|202 | .................... | .................... | .................... | .................... | .................... | .................... |
| AA | 3234 | hCoV-19/Denmark/DCGC-22588/2020\|EPI_ISL_757656\|202 | .................... | .................... | .................... | .................... | .................... | .................... |
| AA | 3235 | hCoV-19/Denmark/DCGC-22589/2020\|EPI_ISL_757657\|202 | .................... | .................... | .................... | .................... | .................... | .................... |
| AA | 3236 | hCoV-19/Denmark/DCGC-22592/2020\|EPI_ISL_757658\|202 | .................... | .................... | .................... | .................... | .................... | .................... |
| AA | 3237 | hCoV-19/Denmark/DCGC-22593/2020\|EPI_ISL_757659\|202 | .................... | .................... | .................... | .................... | .................... | .................... |
| AA | 3238 | hCoV-19/Denmark/DCGC-22595/2020\|EPI_ISL_757660\|202 | .................... | .................... | .................... | .................... | .................... | .................... |
| AA | 3239 | hCoV-19/Denmark/DCGC-22596/2020\|EPI_ISL_757661\|202 | .................... | .................... | .................... | .................... | .................... | .................... |
| AA | 3240 | hCoV-19/Denmark/DCGC-22597/2020\|EPI_ISL_757662\|202 | .................... | .................... | .................... | .................... | .................... | .................... |
| AA | 3241 | hCoV-19/Denmark/DCGC-22601/2020\|EPI_ISL_757663\|202 | .................... | .................... | .................... | .................... | .................... | .................... |
| AA | 3242 | hCoV-19/Denmark/DCGC-22602/2020\|EPI_ISL_757664\|202 | .................... | .................... | .................... | .................... | .................... | .................... |
| AA | 3243 | hCoV-19/Denmark/DCGC-22603/2020\|EPI_ISL_757665\|202 | .................... | .................... | .................... | .................... | .................... | .................... |
| AA | 3244 | hCoV-19/Denmark/DCGC-22604/2020\|EPI_ISL_757666\|202 | .................... | .................... | .................... | .................... | .................... | .................... |
| AA | 3245 | hCoV-19/Denmark/DCGC-22605/2020\|EPI_ISL_757667\|202 | .................... | .................... | .................... | .................... | .................... | .................... |
| AA | 3246 | hCoV-19/Denmark/DCGC-22607/2020\|EPI_ISL_757668\|202 | .................... | .................... | .................... | .................... | .................... | .................... |
| AA | 3247 | hCoV-19/Denmark/DCGC-22610/2020\|EPI_ISL_757669\|202 | .................... | .................... | .................... | .................... | .................... | .................... |
| AA | 3248 | hCoV-19/Denmark/DCGC-22611/2020\|EPI_ISL_757670\|202 | .................... | .................... | .................... | .................... | .................... | .................... |
| AA | 3249 | hCoV-19/Denmark/DCGC-22616/2020\|EPI_ISL_757671\|202 | .................... | .................... | .................... | .................... | .................... | .................... |
| AA | 3250 | hCoV-19/Denmark/DCGC-22618/2020\|EPI_ISL_757673\|202 | .................... | .................... | .................... | .................... | .................... | .................... |
| AA | 3251 | hCoV-19/Denmark/DCGC-22620/2020\|EPI_ISL_757674\|202 | .................... | .................... | .................... | .................... | .................... | .................... |
| AA | 3252 | hCoV-19/Denmark/DCGC-22624/2020\|EPI_ISL_757675\|202 | .................... | .................... | .................... | .................... | .................... | .................... |
| AA | 3253 | hCoV-19/Denmark/DCGC-22625/2020\|EPI_ISL_757676\|202 | .................... | .................... | .................... | .................... | .................... | .................... |
| AA | 3254 | hCoV-19/Denmark/DCGC-22626/2020\|EPI_ISL_757677\|202 | .................... | .................... | .................... | .................... | .................... | .................... |
| AA | 3255 | hCoV-19/Denmark/DCGC-22628/2020\|EPI_ISL_757678\|202 | .................... | .................... | .................... | .................... | .................... | .................... |
| AA | 3256 | hCoV-19/Denmark/DCGC-22631/2020\|EPI_ISL_757679\|202 | .................... | .................... | .................... | .................... | .................... | .................... |
| AA | 3257 | hCoV-19/Denmark/DCGC-20527/2020\|EPI_ISL_758903\|202 | .................... | .................... | .................... | .................... | .................... | .................... |
| AA | 3258 | hCoV-19/Denmark/DCGC-20539/2020\|EPI_ISL_758904\|202 | .................... | .................... | .................... | .................... | .................... | .................... |
| AA | 3259 | hCoV-19/Denmark/DCGC-20543/2020\|EPI_ISL_758905\|202 | .................... | .................... | .................... | .................... | .................... | .................... |
| AA | 3260 | hCoV-19/Denmark/DCGC-20600/2020\|EPI_ISL_758906\|202 | .................... | .................... | .................... | .................... | .................... | .................... |
| AA | 3261 | hCoV-19/Denmark/DCGC-20613/2020\|EPI_ISL_758907\|202 | .................... | .................... | .................... | .................... | .................... | .................... |
| AA | 3262 | hCoV-19/Denmark/DCGC-20620/2020\|EPI_ISL_758908\|202 | .................... | .................... | .................... | .................... | .................... | .................... |
| AA | 3263 | hCoV-19/Denmark/DCGC-20630/2020\|EPI_ISL_758909\|202 | .................... | .................... | .................... | .................... | .................... | .................... |
| AA | 3264 | hCoV-19/Denmark/DCGC-20631/2020\|EPI_ISL_758910\|202 | .................... | .................... | .................... | .................... | .................... | .................... |
| AA | 3265 | hCoV-19/Denmark/DCGC-20632/2020\|EPI_ISL_758911\|202 | .................... | .................... | .................... | .................... | .................... | .................... |
| AA | 3266 | hCoV-19/Denmark/DCGC-20680/2020\|EPI_ISL_758912\|202 | .................... | .................... | .................... | .................... | .................... | .................... |
| AA | 3267 | hCoV-19/Denmark/DCGC-20690/2020\|EPI_ISL_758913\|202 | .................... | .................... | .................... | .................... | .................... | .................... |
| AA | 3268 | hCoV-19/Denmark/DCGC-20741/2020\|EPI_ISL_758914\|202 | .................... | .................... | .................... | .................... | .................... | .................... |
| AA | 3269 | hCoV-19/Denmark/DCGC-20758/2020\|EPI_ISL_758915\|202 | .................... | .................... | .................... | .................... | .................... | .................... |
| AA | 3270 | hCoV-19/Denmark/DCGC-20770/2020\|EPI_ISL_758916\|202 | .................... | .................... | .................... | .................... | .................... | .................... |
| AA | 3271 | hCoV-19/Denmark/DCGC-20771/2020\|EPI_ISL_758917\|202 | .................... | .................... | .................... | .................... | .................... | .................... |
| AA | 3272 | hCoV-19/Denmark/DCGC-20806/2020\|EPI_ISL_758918\|202 | .................... | .................... | .................... | .................... | .................... | .................... |
| AA | 3273 | hCoV-19/Denmark/DCGC-20820/2020\|EPI_ISL_758919\|202 | .................... | .................... | .................... | .................... | .................... | .................... |
| AA | 3274 | hCoV-19/Denmark/DCGC-20834/2020\|EPI_ISL_758920\|202 | .................... | .................... | .................... | .................... | .................... | .................... |
| AA | 3275 | hCoV-19/Denmark/DCGC-20872/2020\|EPI_ISL_758921\|202 | .................... | .................... | .................... | .................... | .................... | .................... |
| AA | 3276 | hCoV-19/Denmark/DCGC-20874/2020\|EPI_ISL_758922\|202 | .................... | .................... | .................... | .................... | .................... | .................... |
| AA | 3277 | hCoV-19/Denmark/DCGC-20876/2020\|EPI_ISL_758923\|202 | .................... | .................... | .................... | .................... | .................... | .................... |
| AA | 3278 | hCoV-19/Denmark/DCGC-20886/2020\|EPI_ISL_758924\|202 | .................... | .................... | .................... | .................... | .................... | .................... |
| AA | 3279 | hCoV-19/Denmark/DCGC-20899/2020\|EPI_ISL_758925\|202 | .................... | .................... | .................... | .................... | .................... | .................... |
| AA | 3280 | hCoV-19/Denmark/DCGC-20925/2020\|EPI_ISL_758926\|202 | .................... | .................... | .................... | .................... | .................... | .................... |
| AA | 3281 | hCoV-19/Denmark/DCGC-20941/2020\|EPI_ISL_758927\|202 | .................... | .................... | .................... | .................... | .................... | .................... |
| AA | 3282 | hCoV-19/Denmark/DCGC-20967/2020\|EPI_ISL_758928\|202 | .................... | .................... | .................... | .................... | .................... | .................... |
| AA | 3283 | hCoV-19/Denmark/DCGC-20973/2020\|EPI_ISL_758929\|202 | .................... | .................... | .................... | .................... | .................... | .................... |

| | | | | | | | | |
|---|---|---|---|---|---|---|---|---|
| AA | 3284 | hCoV-19/Denmark/DCGC-21003/2020\|EPI_ISL_758930\|202 | ···················· | ···················· | ····················· | ···················· | ···················· | ···················· |
| AA | 3285 | hCoV-19/Denmark/DCGC-21004/2020\|EPI_ISL_758931\|202 | ···················· | ···················· | ····················· | ···················· | ···················· | ···················· |
| AA | 3286 | hCoV-19/Denmark/DCGC-21011/2020\|EPI_ISL_758932\|202 | ···················· | ···················· | ····················· | ···················· | ···················· | ···················· |
| AA | 3287 | hCoV-19/Denmark/DCGC-21051/2020\|EPI_ISL_758933\|202 | ···················· | ···················· | ····················· | ···················· | ···················· | ···················· |
| AA | 3288 | hCoV-19/Denmark/DCGC-21074/2020\|EPI_ISL_758934\|202 | ···················· | ···················· | ····················· | ···················· | ···················· | ···················· |
| AA | 3289 | hCoV-19/Denmark/DCGC-21143/2020\|EPI_ISL_758935\|202 | ···················· | ···················· | ····················· | ···················· | ···················· | ···················· |
| AA | 3290 | hCoV-19/Denmark/DCGC-21149/2020\|EPI_ISL_758936\|202 | ···················· | ···················· | ····················· | ···················· | ···················· | ···················· |
| AA | 3291 | hCoV-19/Denmark/DCGC-21212/2020\|EPI_ISL_758937\|202 | ···················· | ···················· | ····················· | ···················· | ···················· | ···················· |
| AA | 3292 | hCoV-19/Denmark/DCGC-21254/2020\|EPI_ISL_758938\|202 | ···················· | ···················· | ····················· | ···················· | ···················· | ···················· |
| AA | 3293 | hCoV-19/Denmark/DCGC-21295/2020\|EPI_ISL_758939\|202 | ···················· | ···················· | ····················· | ···················· | ···················· | ···················· |
| AA | 3294 | hCoV-19/Denmark/DCGC-21297/2020\|EPI_ISL_758940\|202 | ···················· | ···················· | ····················· | ···················· | ···················· | ···················· |
| AA | 3295 | hCoV-19/Denmark/DCGC-21318/2020\|EPI_ISL_758941\|202 | ···················· | ···················· | ····················· | ···················· | ···················· | ···················· |
| AA | 3296 | hCoV-19/Denmark/DCGC-21323/2020\|EPI_ISL_758942\|202 | ···················· | ···················· | ····················· | ···················· | ···················· | ···················· |
| AA | 3297 | hCoV-19/Denmark/DCGC-20986/2020\|EPI_ISL_758943\|202 | ···················· | ···················· | ····················· | ···················· | ···················· | ···················· |
| AA | 3298 | hCoV-19/Denmark/DCGC-20749/2020\|EPI_ISL_758944\|202 | ···················· | ···················· | ····················· | ···················· | ···················· | ···················· |
| AA | 3299 | hCoV-19/Denmark/DCGC-20385/2020\|EPI_ISL_758970\|202 | ···················· | ···················· | ····················· | ···················· | ···················· | ···················· |
| AA | 3300 | hCoV-19/Denmark/DCGC-20718/2020\|EPI_ISL_758971\|202 | ···················· | ···················· | ····················· | ···················· | ···················· | ···················· |
| AA | 3301 | hCoV-19/Denmark/DCGC-20784/2020\|EPI_ISL_758972\|202 | ···················· | ···················· | ····················· | ···················· | ···················· | ···················· |
| AA | 3302 | hCoV-19/Denmark/DCGC-20987/2020\|EPI_ISL_758973\|202 | ···················· | ···················· | ····················· | ···················· | ···················· | ···················· |
| AA | 3303 | hCoV-19/Denmark/DCGC-21156/2020\|EPI_ISL_758974\|202 | ···················· | ···················· | ····················· | ···················· | ···················· | ···················· |
| AA | 3304 | hCoV-19/Denmark/DCGC-21238/2020\|EPI_ISL_758975\|202 | ···················· | ···················· | ····················· | ···················· | ···················· | ···················· |
| AA | 3305 | hCoV-19/Denmark/DCGC-21290/2020\|EPI_ISL_758976\|202 | ···················· | ···················· | ····················· | ···················· | ···················· | ···················· |
| AA | 3306 | hCoV-19/Denmark/DCGC-20362/2020\|EPI_ISL_758977\|202 | ···················· | ···················· | ····················· | ···················· | ···················· | ···················· |
| AA | 3307 | hCoV-19/Denmark/DCGC-20370/2020\|EPI_ISL_758978\|202 | ···················· | ···················· | ····················· | ···················· | ···················· | ···················· |
| AA | 3308 | hCoV-19/Denmark/DCGC-20381/2020\|EPI_ISL_758979\|202 | ···················· | ···················· | ····················· | ···················· | ···················· | ···················· |
| AA | 3309 | hCoV-19/Denmark/DCGC-20387/2020\|EPI_ISL_758980\|202 | ···················· | ···················· | ····················· | ···················· | ···················· | ···················· |
| AA | 3310 | hCoV-19/Denmark/DCGC-20421/2020\|EPI_ISL_758981\|202 | ···················· | ···················· | ····················· | ···················· | ···················· | ···················· |
| AA | 3311 | hCoV-19/Denmark/DCGC-20444/2020\|EPI_ISL_758982\|202 | ···················· | ···················· | ····················· | ···················· | ···················· | ···················· |
| AA | 3312 | hCoV-19/Denmark/DCGC-20459/2020\|EPI_ISL_758983\|202 | ···················· | ···················· | ····················· | ···················· | ···················· | ···················· |
| AA | 3313 | hCoV-19/Denmark/DCGC-20470/2020\|EPI_ISL_758984\|202 | ···················· | ···················· | ····················· | ···················· | ···················· | ···················· |
| AA | 3314 | hCoV-19/Denmark/DCGC-20472/2020\|EPI_ISL_758985\|202 | ···················· | ···················· | ····················· | ···················· | ···················· | ···················· |
| AA | 3315 | hCoV-19/Denmark/DCGC-20479/2020\|EPI_ISL_758986\|202 | ···················· | ···················· | ····················· | ···················· | ···················· | ···················· |
| AA | 3316 | hCoV-19/Denmark/DCGC-20483/2020\|EPI_ISL_758987\|202 | ···················· | ···················· | ····················· | ···················· | ···················· | ···················· |
| AA | 3317 | hCoV-19/Denmark/DCGC-20487/2020\|EPI_ISL_758988\|202 | ···················· | ···················· | ····················· | ···················· | ···················· | ···················· |
| AA | 3318 | hCoV-19/Denmark/DCGC-20489/2020\|EPI_ISL_758989\|202 | ···················· | ···················· | ····················· | ···················· | ··C················· | ···················· |
| AA | 3319 | hCoV-19/Denmark/DCGC-20531/2020\|EPI_ISL_758990\|202 | ···················· | ···················· | ····················· | ···················· | ···················· | ···················· |
| AA | 3320 | hCoV-19/Denmark/DCGC-20540/2020\|EPI_ISL_758991\|202 | ···················· | ···················· | ····················· | ···················· | ···················· | ···················· |
| AA | 3321 | hCoV-19/Denmark/DCGC-20545/2020\|EPI_ISL_758992\|202 | ···················· | ···················· | ····················· | ···················· | ···················· | ···················· |
| AA | 3322 | hCoV-19/Denmark/DCGC-20571/2020\|EPI_ISL_758993\|202 | ···················· | ···················· | ····················· | ···················· | ···················· | ···················· |
| AA | 3323 | hCoV-19/Denmark/DCGC-20599/2020\|EPI_ISL_758994\|202 | ···················· | ···················· | ····················· | ···················· | ···················· | ···················· |
| AA | 3324 | hCoV-19/Denmark/DCGC-20608/2020\|EPI_ISL_758995\|202 | ···················· | ···················· | ····················· | ···················· | ···················· | ···················· |
| AA | 3325 | hCoV-19/Denmark/DCGC-20618/2020\|EPI_ISL_758996\|202 | ···················· | ···················· | ····················· | ···················· | ···T················ | ···················· |
| AA | 3326 | hCoV-19/Denmark/DCGC-20629/2020\|EPI_ISL_758997\|202 | ···················· | ···················· | ····················· | ···················· | ···················· | ···················· |
| AA | 3327 | hCoV-19/Denmark/DCGC-20636/2020\|EPI_ISL_758998\|202 | ···················· | ···················· | ····················· | ···················· | ···················· | ···················· |
| AA | 3328 | hCoV-19/Denmark/DCGC-20660/2020\|EPI_ISL_758999\|202 | ···················· | ···················· | ····················· | ···················· | ···················· | ···················· |
| AA | 3329 | hCoV-19/Denmark/DCGC-17749/2020\|EPI_ISL_749296\|202 | ···················· | ···················· | ····················· | ···················· | ···················· | ···················· |
| AA | 3330 | hCoV-19/Denmark/DCGC-17764/2020\|EPI_ISL_749297\|202 | ···················· | ···················· | ····················· | ···················· | ···················· | ···················· |
| AA | 3331 | hCoV-19/Denmark/DCGC-17779/2020\|EPI_ISL_749298\|202 | ···················· | ···················· | ····················· | ···················· | ···················· | ···················· |
| AA | 3332 | hCoV-19/Denmark/DCGC-17780/2020\|EPI_ISL_749299\|202 | ···················· | ···················· | ····················· | ···················· | ···················· | ···················· |
| AA | 3333 | hCoV-19/Denmark/DCGC-17622/2020\|EPI_ISL_750297\|202 | ···················· | ···················· | ····················· | ···················· | ···················· | ···················· |
| AA | 3334 | hCoV-19/Denmark/DCGC-16758/2020\|EPI_ISL_750298\|202 | ···················· | ···················· | ····················· | ···················· | ···················· | ···················· |
| AA | 3335 | hCoV-19/Denmark/DCGC-16778/2020\|EPI_ISL_750299\|202 | ···················· | ···················· | ····················· | ···················· | ···················· | ···················· |
| AA | 3336 | hCoV-19/USA/CT-CDC-2-3714059/2020\|EPI_ISL_751554\|2 | ···················· | ···················· | ····················· | ···················· | ···················· | ···················· |
| AA | 3337 | hCoV-19/USA/WY-CDC-2-3714468/2020\|EPI_ISL_751555\|2 | ···················· | ···················· | ····················· | ···················· | ···················· | ···················· |
| AA | 3338 | hCoV-19/USA/NV-CDC-2-3714155/2020\|EPI_ISL_751556\|2 | ···················· | ···················· | ····················· | ···················· | ···················· | ···················· |
| AA | 3339 | hCoV-19/USA/NV-CDC-2-3714168/2020\|EPI_ISL_751557\|2 | ···················· | ···················· | ····················· | ···················· | ···················· | ···················· |
| AA | 3340 | hCoV-19/USA/NH-CDC-2-3714170/2020\|EPI_ISL_751558\|2 | ···················· | ···················· | ····················· | ···················· | ···················· | ···················· |
| AA | 3341 | hCoV-19/USA/MD-CDC-2-3713971/2020\|EPI_ISL_751559\|2 | ···················· | ···················· | ····················· | ···················· | ···················· | ···················· |
| AA | 3342 | hCoV-19/USA/NY-CDC-2-3714095/2020\|EPI_ISL_751560\|2 | ···················· | ···················· | ····················· | ···················· | ···················· | ···················· |
| AA | 3343 | hCoV-19/USA/MT-CDC-2-3714162/2020\|EPI_ISL_751561\|2 | ···················· | ···················· | ····················· | ···················· | ···················· | ···················· |
| AA | 3344 | hCoV-19/USA/VA-CDC-2-3714355/2020\|EPI_ISL_751564\|2 | ···················· | ·A·················· | ····················· | ···················· | ···················· | ···················· |
| AA | 3345 | hCoV-19/USA/IA-CDC-2-3714376/2020\|EPI_ISL_751565\|2 | ···················· | ···················· | ····················· | ···················· | ···················· | ···················· |
| AA | 3346 | hCoV-19/USA/WV-CDC-2-3714290/2020\|EPI_ISL_751566\|2 | ···················· | ···················· | ····················· | ···················· | ···················· | ···················· |
| AA | 3347 | hCoV-19/USA/ID-CDC-2-3714453/2020\|EPI_ISL_751567\|2 | ···················· | ···················· | ····················· | ···················· | ···················· | ···················· |
| AA | 3348 | hCoV-19/USA/KY-CDC-2-3714587/2020\|EPI_ISL_751570\|2 | ···················· | ·A·················· | ····················· | ···················· | ···················· | ···················· |
| AA | 3349 | hCoV-19/USA/TX-CDC-2-3714363/2020\|EPI_ISL_751571\|2 | ···················· | ·A·················· | ····················· | ···················· | ···················· | ···················· |
| AA | 3350 | hCoV-19/USA/KS-CDC-2-3714086/2020\|EPI_ISL_751572\|2 | ···················· | ···················· | ····················· | ···················· | ···················· | ···················· |
| AA | 3351 | hCoV-19/USA/MN-CDC-2-3713956/2020\|EPI_ISL_751573\|2 | ···················· | ···················· | ····················· | ···················· | ···················· | ···················· |
| AA | 3352 | hCoV-19/USA/AR-CDC-2-3713957/2020\|EPI_ISL_751574\|2 | ···················· | ···················· | ····················· | ···················· | ···················· | ···················· |
| AA | 3353 | hCoV-19/USA/CO-CDC-2-3713980/2020\|EPI_ISL_751575\|2 | ···················· | ···················· | ····················· | ···················· | ···················· | ···················· |
| AA | 3354 | hCoV-19/USA/MN-CDC-2-3713982/2020\|EPI_ISL_751576\|2 | ···················· | ···················· | ····················· | ···················· | ···················· | ···················· |
| AA | 3355 | hCoV-19/USA/MD-CDC-2-3713985/2020\|EPI_ISL_751577\|2 | ···················· | ···················· | ····················· | ···················· | ···················· | ···················· |
| AA | 3356 | hCoV-19/USA/HI-CDC-2-3714074/2020\|EPI_ISL_751578\|2 | ···················· | ·A·················· | ····················· | ···················· | ···················· | ···················· |
| AA | 3357 | hCoV-19/USA/IN-CDC-2-3714080/2020\|EPI_ISL_751579\|2 | ···················· | ·A·················· | ····················· | ···················· | ···················· | ···················· |
| AA | 3358 | hCoV-19/USA/NY-CDC-2-3714164/2020\|EPI_ISL_751580\|2 | ···················· | ·A·················· | ····················· | ···················· | ···················· | ···················· |
| AA | 3359 | hCoV-19/USA/NY-CDC-2-3714174/2020\|EPI_ISL_751581\|2 | ···················· | ···················· | ····················· | ···················· | ···················· | ···················· |
| AA | 3360 | hCoV-19/USA/NH-CDC-2-3714179/2020\|EPI_ISL_751582\|2 | ···················· | ···················· | ····················· | ···················· | ···················· | ···················· |
| AA | 3361 | hCoV-19/USA/NE-CDC-2-3714192/2020\|EPI_ISL_751583\|2 | ···················· | ···················· | ····················· | ···················· | ···················· | ···················· |
| AA | 3362 | hCoV-19/USA/NH-CDC-2-3714251/2020\|EPI_ISL_751584\|2 | ···················· | ···················· | ····················· | ···················· | ···················· | ···················· |
| AA | 3363 | hCoV-19/USA/NE-CDC-2-3714255/2020\|EPI_ISL_751585\|2 | ···················· | ···················· | ····················· | ···················· | ···················· | ···················· |

| | | | | | | | | |
|---|---|---|---|---|---|---|---|---|
| AA | 3364 | hCoV-19/USA/OH-CDC-2-3714266/2020\|EPI_ISL_751586\|2 | ···················· | ···················· | ····················· | ···················· | ···················· | ···················· |
| AA | 3365 | hCoV-19/USA/NH-CDC-2-3714279/2020\|EPI_ISL_751587\|2 | ···················· | ···················· | ····················· | ···················· | ···················· | ···················· |
| AA | 3366 | hCoV-19/USA/VA-CDC-2-3714379/2020\|EPI_ISL_751588\|2 | ···················· | ···················· | ····················· | ···················· | ···················· | ···················· |
| AA | 3367 | hCoV-19/USA/WV-CDC-2-3714452/2020\|EPI_ISL_751589\|2 | ···················· | ···················· | ····················· | ···················· | ···················· | ···················· |
| AA | 3368 | hCoV-19/USA/ID-CDC-2-3714455/2020\|EPI_ISL_751590\|2 | ···················· | ···················· | ····················· | ···················· | ···················· | ···················· |
| AA | 3369 | hCoV-19/USA/WY-CDC-2-3714467/2020\|EPI_ISL_751591\|2 | ···················· | ···················· | ····················· | ···················· | ···················· | ···················· |
| AA | 3370 | hCoV-19/USA/AK-CDC-2-3714493/2020\|EPI_ISL_751592\|2 | ···················· | ·········A·········· | ····················· | ···················· | ···················· | ···················· |
| AA | 3371 | hCoV-19/USA/AK-CDC-2-3714552/2020\|EPI_ISL_751593\|2 | ···················· | ···················· | ····················· | ···················· | ···················· | ···················· |
| AA | 3372 | hCoV-19/USA/KY-CDC-2-3714554/2020\|EPI_ISL_751594\|2 | ···················· | ···················· | ····················· | ···················· | ···················· | ···················· |
| AA | 3373 | hCoV-19/USA/RI-CDC-2-3714559/2020\|EPI_ISL_751595\|2 | ···················· | ·············A······ | ····················· | ···················· | ···················· | ···················· |
| AA | 3374 | hCoV-19/USA/KY-CDC-2-3714563/2020\|EPI_ISL_751596\|2 | ···················· | ···················· | ····················· | ···················· | ···················· | ···················· |
| AA | 3375 | hCoV-19/USA/AK-CDC-2-3714579/2020\|EPI_ISL_751597\|2 | ···················· | ···················· | ····················· | ···················· | ···················· | ···················· |
| AA | 3376 | hCoV-19/USA/AK-CDC-2-3714583/2020\|EPI_ISL_751598\|2 | ···················· | ···················· | ····················· | ···················· | ···················· | ···················· |
| AA | 3377 | hCoV-19/USA/IN-CDC-2-3713959/2020\|EPI_ISL_751599\|2 | ···················· | ···················· | ····················· | ···················· | ···················· | ···················· |
| AA | 3378 | hCoV-19/USA/HI-H200241/2020\|EPI_ISL_752800\|2020-08 | ···················· | ·A·················· | ····················· | ···················· | ···················· | ···················· |
| AA | 3379 | hCoV-19/USA/HI-H200242/2020\|EPI_ISL_752801\|2020-08 | ···················· | ·A·················· | ····················· | ···················· | ···················· | ···················· |
| AA | 3380 | hCoV-19/USA/HI-H200243/2020\|EPI_ISL_752802\|2020-08 | ···················· | ·A·················· | ····················· | ···················· | ···················· | ···················· |
| AA | 3381 | hCoV-19/USA/HI-H200244/2020\|EPI_ISL_752803\|2020-08 | ···················· | ·A·················· | ····················· | ···················· | ···················· | ···················· |
| AA | 3382 | hCoV-19/USA/HI-H200245/2020\|EPI_ISL_752804\|2020-08 | ···················· | ·A·················· | ····················· | ···················· | ···················· | ···················· |
| AA | 3383 | hCoV-19/USA/HI-H200246/2020\|EPI_ISL_752805\|2020-08 | ···················· | ·A·················· | ····················· | ···················· | ···················· | ···················· |
| AA | 3384 | hCoV-19/USA/HI-H200247/2020\|EPI_ISL_752806\|2020-08 | ···················· | ·A·················· | ····················· | ···················· | ···················· | ···················· |
| AA | 3385 | hCoV-19/USA/HI-H200248/2020\|EPI_ISL_752807\|2020-08 | ···················· | ·A·················· | ····················· | ···················· | ···················· | ···················· |
| AA | 3386 | hCoV-19/USA/HI-H200249/2020\|EPI_ISL_752808\|2020-08 | ···················· | ·A·················· | ····················· | ···················· | ···················· | ···················· |
| AA | 3387 | hCoV-19/USA/HI-H200250/2020\|EPI_ISL_752809\|2020-08 | ···················· | ·A·················· | ····················· | ···················· | ···················· | ···················· |
| AA | 3388 | hCoV-19/USA/HI-H200251/2020\|EPI_ISL_752810\|2020-08 | ···················· | ·A·················· | ····················· | ···················· | ···················· | ···················· |
| AA | 3389 | hCoV-19/USA/HI-H200252/2020\|EPI_ISL_752811\|2020-08 | ···················· | ·A·················· | ····················· | ···················· | ···················· | ···················· |
| AA | 3390 | hCoV-19/USA/HI-H200253/2020\|EPI_ISL_752812\|2020-08 | ···················· | ·A·················· | ····················· | ···················· | ···················· | ···················· |
| AA | 3391 | hCoV-19/USA/HI-H200254/2020\|EPI_ISL_752813\|2020-08 | ···················· | ·A·················· | ····················· | ···················· | ···················· | ···················· |
| AA | 3392 | hCoV-19/USA/HI-H200255/2020\|EPI_ISL_752814\|2020-08 | ···················· | ·A·················· | ····················· | ···················· | ···················· | ···················· |
| AA | 3393 | hCoV-19/USA/HI-H200256/2020\|EPI_ISL_752815\|2020-08 | ···················· | ·A·················· | ····················· | ···················· | ···················· | ···················· |
| AA | 3394 | hCoV-19/USA/HI-H200257/2020\|EPI_ISL_752816\|2020-08 | ···················· | ·A·················· | ····················· | ···················· | ···················· | ···················· |
| AA | 3395 | hCoV-19/USA/HI-H200258/2020\|EPI_ISL_752817\|2020-08 | ···················· | ·A·················· | ····················· | ···················· | ···················· | ···················· |
| AA | 3396 | hCoV-19/USA/HI-H200259/2020\|EPI_ISL_752818\|2020-08 | ···················· | ·A·················· | ····················· | ···················· | ···················· | ···················· |
| AA | 3397 | hCoV-19/USA/HI-H200260/2020\|EPI_ISL_752819\|2020-08 | ···················· | ·A·················· | ····················· | ···················· | ···················· | ···················· |
| AA | 3398 | hCoV-19/USA/HI-H200261/2020\|EPI_ISL_752820\|2020-08 | ···················· | ·A·················· | ····················· | ···················· | ···················· | ···················· |
| AA | 3399 | hCoV-19/USA/HI-H200262/2020\|EPI_ISL_752821\|2020-08 | ···················· | ·A·················· | ····················· | ···················· | ···················· | ···················· |
| AA | 3400 | hCoV-19/USA/HI-H200263/2020\|EPI_ISL_752822\|2020-08 | ···················· | ·A·················· | ····················· | ···················· | ···················· | ···················· |
| AA | 3401 | hCoV-19/USA/HI-H200264/2020\|EPI_ISL_752823\|2020-08 | ···················· | ·A·················· | ····················· | ···················· | ···················· | ···················· |
| AA | 3402 | hCoV-19/USA/HI-H200266/2020\|EPI_ISL_752825\|2020-08 | ···················· | ·A·················· | ····················· | ···················· | ···················· | ···················· |
| AA | 3403 | hCoV-19/USA/HI-H200267/2020\|EPI_ISL_752826\|2020-09 | ···················· | ·A·················· | ····················· | ···················· | ···················· | ···················· |
| AA | 3404 | hCoV-19/USA/HI-H200268/2020\|EPI_ISL_752827\|2020-09 | ···················· | ·A·················· | ····················· | ···················· | ···················· | ···················· |
| AA | 3405 | hCoV-19/USA/HI-H200269/2020\|EPI_ISL_752828\|2020-09 | ···················· | ·A·················· | ····················· | ···················· | ···················· | ···················· |
| AA | 3406 | hCoV-19/USA/HI-H200270/2020\|EPI_ISL_752829\|2020-09 | ···················· | ·A·················· | ····················· | ···················· | ···················· | ···················· |
| AA | 3407 | hCoV-19/USA/HI-H200271/2020\|EPI_ISL_752830\|2020-09 | ···················· | ·A·················· | ····················· | ···················· | ···················· | ···················· |
| AA | 3408 | hCoV-19/USA/HI-H200272/2020\|EPI_ISL_752831\|2020-09 | ···················· | ·A·················· | ····················· | ···················· | ···················· | ···················· |
| AA | 3409 | hCoV-19/USA/HI-H200273/2020\|EPI_ISL_752832\|2020-09 | ···················· | ·A·················· | ····················· | ···················· | ···················· | ···················· |
| AA | 3410 | hCoV-19/USA/HI-H200274/2020\|EPI_ISL_752833\|2020-09 | ···················· | ·A·················· | ····················· | ···················· | ···················· | ···················· |
| AA | 3411 | hCoV-19/USA/HI-H200275/2020\|EPI_ISL_752834\|2020-09 | ···················· | ·A·················· | ····················· | ···················· | ···················· | ···················· |
| AA | 3412 | hCoV-19/USA/HI-H200276/2020\|EPI_ISL_752835\|2020-09 | ···················· | ·A·················· | ····················· | ···················· | ···················· | ···················· |
| AA | 3413 | hCoV-19/USA/HI-H200277/2020\|EPI_ISL_752836\|2020-09 | ···················· | ·A·················· | ····················· | ···················· | ···················· | ···················· |
| AA | 3414 | hCoV-19/USA/HI-H200278/2020\|EPI_ISL_752837\|2020-09 | ···················· | ·A·················· | ····················· | ···················· | ···················· | ···················· |
| AA | 3415 | hCoV-19/USA/HI-H200279/2020\|EPI_ISL_752838\|2020-09 | ···················· | ·A·················· | ····················· | ···················· | ···················· | ···················· |
| AA | 3416 | hCoV-19/USA/HI-H200280/2020\|EPI_ISL_752839\|2020-09 | ···················· | ·A·················· | ····················· | ···················· | ···················· | ···················· |
| AA | 3417 | hCoV-19/USA/HI-H200281/2020\|EPI_ISL_752840\|2020-09 | ···················· | ·A·················· | ····················· | ···················· | ···················· | ···················· |
| AA | 3418 | hCoV-19/USA/HI-H200282/2020\|EPI_ISL_752841\|2020-09 | ···················· | ·A·················· | ····················· | ···················· | ···················· | ···················· |
| AA | 3419 | hCoV-19/USA/HI-H200283/2020\|EPI_ISL_752842\|2020-09 | ···················· | ·A·················· | ····················· | ···················· | ···················· | ···················· |
| AA | 3420 | hCoV-19/USA/HI-H200284/2020\|EPI_ISL_752843\|2020-09 | ···················· | ·A·················· | ····················· | ···················· | ···················· | ···················· |
| AA | 3421 | hCoV-19/USA/HI-H200285/2020\|EPI_ISL_752844\|2020-07 | ···················· | ···················· | ····················· | ···················· | ···················· | ···················· |
| AA | 3422 | hCoV-19/USA/HI-H200286/2020\|EPI_ISL_752845\|2020-03 | ···················· | ·A·················· | ····················· | ···················· | ···················· | ···················· |
| AA | 3423 | hCoV-19/USA/HI-H200287/2020\|EPI_ISL_752846\|2020-08 | ···················· | ·A·················· | ····················· | ···················· | ···················· | ···················· |
| AA | 3424 | hCoV-19/USA/HI-H200288/2020\|EPI_ISL_752847\|2020-08 | ···················· | ·A·················· | ····················· | ···················· | ···················· | ···················· |
| AA | 3425 | hCoV-19/USA/HI-H200289/2020\|EPI_ISL_752848\|2020-08 | ···················· | ·A·················· | ····················· | ···················· | ···················· | ···················· |
| AA | 3426 | hCoV-19/USA/HI-H200290/2020\|EPI_ISL_752849\|2020-08 | ···················· | ·A·················· | ····················· | ···················· | ···················· | ···················· |
| AA | 3427 | hCoV-19/USA/HI-H200291/2020\|EPI_ISL_752850\|2020-08 | ···················· | ···················· | ····················· | ···················· | ···················· | ···················· |
| AA | 3428 | hCoV-19/USA/HI-H200292/2020\|EPI_ISL_752851\|2020-08 | ···················· | ·A·················· | ····················· | ···················· | ···················· | ···················· |
| AA | 3429 | hCoV-19/USA/HI-H200293/2020\|EPI_ISL_752852\|2020-08 | ···················· | ·A·················· | ····················· | ···················· | ···················· | ···················· |
| AA | 3430 | hCoV-19/USA/HI-H200294/2020\|EPI_ISL_752853\|2020-08 | ···················· | ·A·················· | ····················· | ···················· | ···················· | ···················· |
| AA | 3431 | hCoV-19/USA/HI-H200295/2020\|EPI_ISL_752854\|2020-08 | ···················· | ·A·················· | ····················· | ···················· | ···················· | ···················· |
| AA | 3432 | hCoV-19/USA/HI-H200296/2020\|EPI_ISL_752855\|2020-08 | ···················· | ·A·················· | ····················· | ···················· | ···················· | ···················· |
| AA | 3433 | hCoV-19/USA/HI-H200297/2020\|EPI_ISL_752856\|2020-08 | ···················· | ·A·················· | ····················· | ···················· | ···················· | ···················· |
| AA | 3434 | hCoV-19/USA/HI-H200298/2020\|EPI_ISL_752857\|2020-08 | ···················· | ·A·················· | ····················· | ···················· | ···················· | ···················· |
| AA | 3435 | hCoV-19/USA/HI-H200299/2020\|EPI_ISL_752858\|2020-08 | ···················· | ·A·················· | ····················· | ···················· | ···················· | ···················· |
| AA | 3436 | hCoV-19/USA/HI-H200300/2020\|EPI_ISL_752859\|2020-08 | ···················· | ·A·················· | ····················· | ···················· | ···················· | ···················· |
| AA | 3437 | hCoV-19/USA/HI-H200301/2020\|EPI_ISL_752860\|2020-08 | ···················· | ·A·················· | ····················· | ···················· | ···················· | ···················· |
| AA | 3438 | hCoV-19/USA/HI-H200302/2020\|EPI_ISL_752861\|2020-08 | ···················· | ·A·················· | ····················· | ···················· | ···················· | ···················· |
| AA | 3439 | hCoV-19/USA/HI-H200303/2020\|EPI_ISL_752862\|2020-08 | ···················· | ·A·················· | ····················· | ···················· | ···················· | ···················· |
| AA | 3440 | hCoV-19/USA/HI-H200304/2020\|EPI_ISL_752863\|2020-08 | ···················· | ·A·················· | ····················· | ···················· | ···················· | ···················· |
| AA | 3441 | hCoV-19/USA/HI-H200305/2020\|EPI_ISL_752864\|2020-08 | ···················· | ·A·················· | ····················· | ···················· | ···················· | ···················· |
| AA | 3442 | hCoV-19/USA/HI-H200306/2020\|EPI_ISL_752865\|2020-08 | ···················· | ·A·················· | ····················· | ···················· | ···················· | ···················· |
| AA | 3443 | hCoV-19/USA/HI-H200307/2020\|EPI_ISL_752866\|2020-08 | ···················· | ·A·················· | ····················· | ···················· | ···················· | ···················· |

| | | | | | | | | |
|---|---|---|---|---|---|---|---|---|
| AA | 3444 | hCoV-19/USA/HI-H200308/2020\|EPI_ISL_752867\|2020-08 | .................... | .A.................. | .................... | .................... | .................... | .................... |
| AA | 3445 | hCoV-19/USA/HI-H200309/2020\|EPI_ISL_752868\|2020-08 | .................... | .A.................. | .................... | .................... | .................... | .................... |
| AA | 3446 | hCoV-19/USA/HI-H200310/2020\|EPI_ISL_752869\|2020-08 | .................... | .A.................. | .................... | .................... | .................... | .................... |
| AA | 3447 | hCoV-19/USA/HI-H200311/2020\|EPI_ISL_752870\|2020-08 | .................... | .A.................. | .................... | .................... | .................... | .................... |
| AA | 3448 | hCoV-19/USA/HI-H200312/2020\|EPI_ISL_752871\|2020-08 | .................... | .A.................. | .................... | .................... | .................... | .................... |
| AA | 3449 | hCoV-19/USA/HI-H200313/2020\|EPI_ISL_752872\|2020-08 | .................... | .A.................. | .................... | .................... | .................... | .................... |
| AA | 3450 | hCoV-19/USA/HI-H200314/2020\|EPI_ISL_752873\|2020-08 | .................... | .A.................. | .................... | .................... | .................... | .................... |
| AA | 3451 | hCoV-19/USA/HI-H200315/2020\|EPI_ISL_752874\|2020-08 | .................... | .A.................. | .................... | .................... | .................... | .................... |
| AA | 3452 | hCoV-19/USA/HI-H200316/2020\|EPI_ISL_752875\|2020-08 | .................... | .A.................. | .................... | .................... | .................... | .................... |
| AA | 3453 | hCoV-19/USA/HI-H200317/2020\|EPI_ISL_752876\|2020-08 | .................... | .A.................. | .................... | .................... | .................... | .................... |
| AA | 3454 | hCoV-19/USA/HI-H200318/2020\|EPI_ISL_752877\|2020-08 | .................... | .A.................. | .................... | .................... | .................... | .................... |
| AA | 3455 | hCoV-19/USA/HI-H200319/2020\|EPI_ISL_752878\|2020-08 | .................... | .A.................. | .................... | .................... | .................... | .................... |
| AA | 3456 | hCoV-19/USA/HI-H200320/2020\|EPI_ISL_752879\|2020-08 | .................... | .................... | .................... | .................... | .................... | .................... |
| AA | 3457 | hCoV-19/USA/HI-H200322/2020\|EPI_ISL_752880\|2020-08 | .................... | .A.................. | .................... | .................... | .................... | .................... |
| AA | 3458 | hCoV-19/USA/HI-H200323/2020\|EPI_ISL_752881\|2020-08 | .................... | .A.................. | .................... | .................... | .................... | .................... |
| AA | 3459 | hCoV-19/USA/HI-H200324/2020\|EPI_ISL_752882\|2020-08 | .................... | .A.................. | .................... | .................... | .................... | .................... |
| AA | 3460 | hCoV-19/USA/HI-H200325/2020\|EPI_ISL_752883\|2020-08 | .................... | .A.................. | .................... | .................... | .................... | .................... |
| AA | 3461 | hCoV-19/USA/HI-H200326/2020\|EPI_ISL_752884\|2020-08 | .................... | .A.................. | .................... | .................... | .................... | .................... |
| AA | 3462 | hCoV-19/USA/HI-H200327/2020\|EPI_ISL_752885\|2020-08 | .................... | .A.................. | .................... | .................... | .................... | .................... |
| AA | 3463 | hCoV-19/USA/HI-H200328/2020\|EPI_ISL_752886\|2020-08 | .................... | .A.................. | .................... | .................... | .................... | .................... |
| AA | 3464 | hCoV-19/USA/HI-H200329/2020\|EPI_ISL_752887\|2020-08 | .................... | .A.................. | .................... | .................... | .................... | .................... |
| AA | 3465 | hCoV-19/USA/HI-H200330/2020\|EPI_ISL_752888\|2020-08 | .................... | .A.................. | .................... | .................... | .................... | .................... |
| AA | 3466 | hCoV-19/USA/HI-H200331/2020\|EPI_ISL_752889\|2020-08 | .................... | .A.................. | .................... | .................... | .................... | .................... |
| AA | 3467 | hCoV-19/USA/HI-H200332/2020\|EPI_ISL_752890\|2020-08 | .................... | .A.................. | .................... | .................... | .................... | .................... |
| AA | 3468 | hCoV-19/USA/HI-H200333/2020\|EPI_ISL_752891\|2020-08 | .................... | .A.................. | .................... | .................... | .................... | .................... |
| AA | 3469 | hCoV-19/USA/HI-H200334/2020\|EPI_ISL_752892\|2020-08 | .................... | .A.................. | .................... | .................... | .................... | .................... |
| AA | 3470 | hCoV-19/USA/HI-H200335/2020\|EPI_ISL_752893\|2020-08 | .................... | .A.................. | .................... | .................... | .................... | .................... |
| AA | 3471 | hCoV-19/USA/HI-H200336/2020\|EPI_ISL_752894\|2020-08 | .................... | .A.................. | .................... | .................... | .................... | .................... |
| AA | 3472 | hCoV-19/USA/HI-H200337/2020\|EPI_ISL_752895\|2020-08 | .................... | .A.................. | .................... | .................... | .................... | .................... |
| AA | 3473 | hCoV-19/USA/HI-H200339/2020\|EPI_ISL_752896\|2020-08 | .................... | .A.................. | .................... | .................... | .................... | .................... |
| AA | 3474 | hCoV-19/USA/HI-H200340/2020\|EPI_ISL_752897\|2020-08 | .................... | .A.................. | .................... | .................... | .................... | .................... |
| AA | 3475 | hCoV-19/USA/HI-H200342/2020\|EPI_ISL_752898\|2020-08 | .................... | .A.................. | .................... | .................... | .................... | .................... |
| AA | 3476 | hCoV-19/USA/HI-H200343/2020\|EPI_ISL_752899\|2020-08 | .................... | .A.................. | .................... | .................... | .................... | .................... |
| AA | 3477 | hCoV-19/USA/CA-CDPH291/2020\|EPI_ISL_755030\|2020-05 | .................... | .................... | .................... | .................... | .................... | .................... |
| AA | 3478 | hCoV-19/USA/CA-CDPH158/2020\|EPI_ISL_755031\|2020-09 | .................... | .................... | .................... | .................... | .................... | .................... |
| AA | 3479 | hCoV-19/USA/CA-CDPH159/2020\|EPI_ISL_755032\|2020-09 | .................... | .................... | .................... | .................... | .................... | .................... |
| AA | 3480 | hCoV-19/USA/CA-CDPH045/2020\|EPI_ISL_755033\|2020-05 | .................... | .................... | .................... | .................... | .................... | .................... |
| AA | 3481 | hCoV-19/USA/CA-CDPH058/2020\|EPI_ISL_755034\|2020-06 | .................... | .................... | .................... | .................... | .................... | .................... |
| AA | 3482 | hCoV-19/USA/CA-CDPH059/2020\|EPI_ISL_755035\|2020-06 | .................... | .................... | .................... | .................... | .................... | .................... |
| AA | 3483 | hCoV-19/USA/CA-CDPH064/2020\|EPI_ISL_755036\|2020-03 | .................... | .................... | .................... | .................... | .................... | .................... |
| AA | 3484 | hCoV-19/USA/CA-CDPH065/2020\|EPI_ISL_755037\|2020-03 | .................... | .................... | .................... | .................... | .................... | .................... |
| AA | 3485 | hCoV-19/USA/CA-CDPH066/2020\|EPI_ISL_755038\|2020-04 | .................... | .................... | .................... | .................... | .................... | .................... |
| AA | 3486 | hCoV-19/USA/CA-CDPH067/2020\|EPI_ISL_755039\|2020-04 | .................... | .................... | .................... | .................... | .................... | .................... |
| AA | 3487 | hCoV-19/USA/CA-CDPH230/2020\|EPI_ISL_755080\|2020-11 | .................... | .................... | .................... | .................... | .................... | .................... |
| AA | 3488 | hCoV-19/USA/CA-CDPH234/2020\|EPI_ISL_755081\|2020-06 | .................... | .................... | .................... | .................... | .................... | .................... |
| AA | 3489 | hCoV-19/USA/CA-CDPH236/2020\|EPI_ISL_755082\|2020-11 | .................... | .................... | .................... | .................... | .................... | .................... |
| AA | 3490 | hCoV-19/USA/CA-CDPH237/2020\|EPI_ISL_755083\|2020-11 | .................... | .................... | .................... | .................... | .................... | .................... |
| AA | 3491 | hCoV-19/USA/CA-CDPH239/2020\|EPI_ISL_755084\|2020-11 | .................... | .................... | .................... | .................... | .................... | .................... |
| AA | 3492 | hCoV-19/USA/CA-CDPH240/2020\|EPI_ISL_755085\|2020-11 | .................... | .................... | .................... | .................... | .................... | .................... |
| AA | 3493 | hCoV-19/USA/CA-CDPH241/2020\|EPI_ISL_755086\|2020-11 | .................... | .................... | .................... | .................... | .................... | .................... |
| AA | 3494 | hCoV-19/USA/CA-CDPH244/2020\|EPI_ISL_755087\|2020-11 | .................... | .................... | .................... | .................... | .................... | .................... |
| AA | 3495 | hCoV-19/USA/CA-CDPH248/2020\|EPI_ISL_755088\|2020-11 | .................... | .................... | .................... | .................... | .................... | .................... |
| AA | 3496 | hCoV-19/USA/CA-CDPH251/2020\|EPI_ISL_755089\|2020-11 | .................... | .................... | .................... | .................... | .................... | .................... |
| AA | 3497 | hCoV-19/USA/CA-CDPH254/2020\|EPI_ISL_755090\|2020-11 | .................... | .................... | .................... | .................... | .................... | .................... |
| AA | 3498 | hCoV-19/USA/CA-CDPH255/2020\|EPI_ISL_755091\|2020-11 | .................... | .................... | .................... | .................... | .................... | .................... |
| AA | 3499 | hCoV-19/USA/CA-CDPH258/2020\|EPI_ISL_755092\|2020-11 | .................... | .................... | .................... | .................... | .................... | .................... |
| AA | 3500 | hCoV-19/USA/CA-CDPH266/2020\|EPI_ISL_755093\|2020-11 | .................... | .................... | .................... | .................... | .................... | .................... |
| AA | 3501 | hCoV-19/USA/CA-CDPH267/2020\|EPI_ISL_755094\|2020-11 | .................... | .................... | .................... | .................... | .................... | .................... |
| AA | 3502 | hCoV-19/USA/CA-CDPH268/2020\|EPI_ISL_755095\|2020-11 | .................... | .................... | .................... | .................... | .................... | .................... |
| AA | 3503 | hCoV-19/USA/CA-CDPH269/2020\|EPI_ISL_755096\|2020-11 | .................... | .................... | .................... | .................... | .................... | .................... |
| AA | 3504 | hCoV-19/USA/CA-CDPH270/2020\|EPI_ISL_755097\|2020-11 | .................... | .................... | .................... | .................... | .................... | .................... |
| AA | 3505 | hCoV-19/USA/CA-CDPH271/2020\|EPI_ISL_755098\|2020-11 | .................... | .................... | .................... | .................... | .................... | .................... |
| AA | 3506 | hCoV-19/USA/CA-CDPH272/2020\|EPI_ISL_755099\|2020-11 | .................... | .................... | .................... | .................... | .................... | .................... |
| AA | 3507 | hCoV-19/USA/CA-CDPH273/2020\|EPI_ISL_755100\|2020-11 | .................... | .................... | .................... | .................... | .................... | .................... |
| AA | 3508 | hCoV-19/USA/CA-CDPH275/2020\|EPI_ISL_755101\|2020-11 | .................... | .................... | .................... | .................... | .................... | .................... |
| AA | 3509 | hCoV-19/USA/CA-CDPH276/2020\|EPI_ISL_755102\|2020-11 | .................... | .................... | .................... | .................... | .................... | .................... |
| AA | 3510 | hCoV-19/USA/CA-CDPH277/2020\|EPI_ISL_755103\|2020-11 | .................... | .................... | .................... | .................... | .................... | .................... |
| AA | 3511 | hCoV-19/USA/CA-CDPH278/2020\|EPI_ISL_755104\|2020-11 | .................... | .................... | .................... | .................... | .................... | .................... |
| AA | 3512 | hCoV-19/USA/CA-CDPH279/2020\|EPI_ISL_755105\|2020-11 | .................... | .................... | .................... | .................... | .................... | .................... |
| AA | 3513 | hCoV-19/USA/CA-CDPH280/2020\|EPI_ISL_755106\|2020-11 | .................... | .................... | .................... | .................... | .................... | .................... |
| AA | 3514 | hCoV-19/USA/CA-CDPH282/2020\|EPI_ISL_755107\|2020-11 | .................... | .................... | .................... | .................... | .................... | .................... |
| AA | 3515 | hCoV-19/USA/CA-CDPH283/2020\|EPI_ISL_755108\|2020-11 | .................... | .................... | .................... | .................... | .................... | .................... |
| AA | 3516 | hCoV-19/USA/CA-CDPH287/2020\|EPI_ISL_755109\|2020-11 | .................... | .................... | .................... | .................... | .................... | .................... |
| AA | 3517 | hCoV-19/USA/CA-CDPH288/2020\|EPI_ISL_755110\|2020-11 | .................... | .................... | .................... | .................... | .................... | .................... |
| AA | 3518 | hCoV-19/USA/CA-CDPH290/2020\|EPI_ISL_755111\|2020-11 | .................... | .................... | .................... | .................... | .................... | .................... |
| AA | 3519 | hCoV-19/USA/CA-CDPH293/2020\|EPI_ISL_755112\|2020-12 | .................... | .................... | .................... | .................... | .................... | .................... |
| AA | 3520 | hCoV-19/USA/CA-CDPH295/2020\|EPI_ISL_755113\|2020-12 | .................... | .................... | .................... | .................... | .................... | .................... |
| AA | 3521 | hCoV-19/USA/CA-CDPH296/2020\|EPI_ISL_755114\|2020-12 | .................... | .................... | .................... | .................... | .................... | .................... |
| AA | 3522 | hCoV-19/USA/CA-SEARCH-5343/2020\|EPI_ISL_755160\|202 | .................... | .................... | .................... | .................... | .................... | .................... |
| AA | 3523 | hCoV-19/USA/CA-SEARCH-5344/2020\|EPI_ISL_755161\|202 | .................... | .................... | .................... | .................... | .................... | .................... |

| | | | | | | | |
|---|---|---|---|---|---|---|---|
| AA | 3524 hCoV-19/USA/CA-SEARCH-5347/2020\|EPI_ISL_755162\|202 | ···················· | ···················· | ···················· | ···················· | ···················· | ···················· |
| AA | 3525 hCoV-19/USA/CA-SEARCH-5348/2020\|EPI_ISL_755163\|202 | ···················· | ···················· | ···················· | ···················· | ···················· | ···················· |
| AA | 3526 hCoV-19/USA/CA-SEARCH-5349/2020\|EPI_ISL_755164\|202 | ···················· | ···················· | ···················· | ···················· | ···················· | ···················· |
| AA | 3527 hCoV-19/USA/CA-SEARCH-5350/2020\|EPI_ISL_755165\|202 | ···················· | ·A·················· | ···················· | ···················· | ···················· | ···················· |
| AA | 3528 hCoV-19/USA/CA-SEARCH-5351/2020\|EPI_ISL_755166\|202 | ···················· | ·A·················· | ···················· | ···················· | ···················· | ···················· |
| AA | 3529 hCoV-19/USA/CA-SEARCH-5352/2020\|EPI_ISL_755167\|202 | ···················· | ·A·················· | ···················· | ···················· | ···················· | ···················· |
| AA | 3530 hCoV-19/USA/CA-SEARCH-5353/2020\|EPI_ISL_755168\|202 | ···················· | ···················· | ···················· | ···················· | ···················· | ···················· |
| AA | 3531 hCoV-19/USA/CA-SEARCH-5356/2020\|EPI_ISL_755169\|202 | ···················· | ·A·················· | ···················· | ···················· | ···················· | ···················· |
| AA | 3532 hCoV-19/USA/CA-SEARCH-5357/2020\|EPI_ISL_755170\|202 | ···················· | ···················· | ···················· | ···················· | ···················· | ···················· |
| AA | 3533 hCoV-19/USA/CA-SEARCH-5365/2020\|EPI_ISL_755177\|202 | ···················· | ···················· | ···················· | ···················· | ···················· | ···················· |
| AA | 3534 hCoV-19/USA/CA-SEARCH-5366/2020\|EPI_ISL_755178\|202 | ···········T········ | ···················· | ···················· | ···················· | ···················· | ···················· |
| AA | 3535 hCoV-19/USA/CA-SEARCH-5367/2020\|EPI_ISL_755179\|202 | ···················· | ···················· | ···················· | ···················· | ···················· | ···················· |
| AA | 3536 hCoV-19/USA/CA-IGI-0805/2020\|EPI_ISL_756341\|2020-0 | ···················· | ···················· | ···················· | ···················· | ···················· | ···················· |
| AA | 3537 hCoV-19/USA/CA-IGI-0827/2020\|EPI_ISL_756351\|2020-0 | ···················· | ···················· | ···················· | ···················· | ···················· | ···················· |
| AA | 3538 hCoV-19/USA/CA-IGI-0829/2020\|EPI_ISL_756352\|2020-0 | ···················· | ···················· | ···················· | ···················· | ···················· | ···················· |
| AA | 3539 hCoV-19/USA/CA-IGI-0831/2020\|EPI_ISL_756353\|2020-0 | ···················· | ···················· | ···················· | ···················· | ···················· | ···················· |
| AA | 3540 hCoV-19/USA/CA-IGI-0832/2020\|EPI_ISL_756354\|2020-0 | ···················· | ···················· | ···················· | ···················· | ···················· | ···················· |
| AA | 3541 hCoV-19/USA/CA-IGI-0834_H06/2020\|EPI_ISL_756355\|20 | ···················· | ···················· | ···················· | ···················· | ···················· | ···················· |
| AA | 3542 hCoV-19/USA/CA-IGI-0837/2020\|EPI_ISL_756356\|2020-0 | ···················· | ···················· | ···················· | ···················· | ···················· | ···················· |
| AA | 3543 hCoV-19/Indonesia/SU-NIHRDC-0045870/2020\|EPI_ISL_7 | ···················· | ···················· | ···················· | ···················· | ···················· | ··A················· |
| AA | 3544 hCoV-19/England/ALDP-C8B61E/2020\|EPI_ISL_756402\|20 | ···················· | ···················· | ···················· | ···················· | ···················· | ···················· |
| AA | 3545 hCoV-19/England/ALDP-C8B54E/2020\|EPI_ISL_756407\|20 | ···················· | ···················· | ···················· | ···················· | ···················· | ···················· |
| AA | 3546 hCoV-19/England/ALDP-C8AC2C/2020\|EPI_ISL_756408\|20 | ···················· | ···················· | ···················· | ···················· | ···················· | ···················· |
| AA | 3547 hCoV-19/England/CAMC-C85957/2020\|EPI_ISL_756409\|20 | ···················· | ·A·················· | ···················· | ···················· | ···················· | ···················· |
| AA | 3548 hCoV-19/England/ALDP-C88435/2020\|EPI_ISL_756414\|20 | ···················· | ···················· | ···················· | ···················· | ···················· | ···················· |
| AA | 3549 hCoV-19/England/ALDP-C8B748/2020\|EPI_ISL_756423\|20 | ···················· | ···················· | ···················· | ···················· | ···················· | ···················· |
| AA | 3550 hCoV-19/England/CAMC-C86631/2020\|EPI_ISL_756429\|20 | ···················· | ···················· | ···················· | ···················· | ···················· | ···················· |
| AA | 3551 hCoV-19/England/ALDP-C8B450/2020\|EPI_ISL_756435\|20 | ···················· | ···················· | ···················· | ···················· | ·····T·············· | ···················· |
| AA | 3552 hCoV-19/England/CAMC-C84554/2020\|EPI_ISL_756438\|20 | ···················· | ···················· | ···················· | ···················· | ···················· | ···················· |
| AA | 3553 hCoV-19/England/ALDP-C8B414/2020\|EPI_ISL_756439\|20 | ···················· | ···················· | ···················· | ···················· | ···················· | ···················· |
| AA | 3554 hCoV-19/England/CAMC-C85456/2020\|EPI_ISL_756444\|20 | ···················· | ···················· | ···················· | ···················· | ···················· | ···················· |
| AA | 3555 hCoV-19/England/ALDP-C8A22A/2020\|EPI_ISL_756445\|20 | ···················· | ···················· | ···················· | ···················· | ···················· | ···················· |
| AA | 3556 hCoV-19/England/CAMC-C85A72/2020\|EPI_ISL_756448\|20 | ···················· | ···················· | ···················· | ···················· | ···················· | ···················· |
| AA | 3557 hCoV-19/England/ALDP-C8B669/2020\|EPI_ISL_756449\|20 | ···················· | ···················· | ···················· | ···················· | ···················· | ···················· |
| AA | 3558 hCoV-19/England/CAMC-C859B1/2020\|EPI_ISL_756490\|20 | ···················· | ···················· | ···················· | ···················· | ···················· | ···················· |
| AA | 3559 hCoV-19/England/CAMC-C83FDF/2020\|EPI_ISL_756491\|20 | ···················· | ···················· | ···················· | ···················· | ···················· | ···················· |
| AA | 3560 hCoV-19/England/CAMC-C84E86/2020\|EPI_ISL_756492\|20 | ···················· | ···················· | ···················· | ···················· | ·····T·············· | ···················· |
| AA | 3561 hCoV-19/England/ALDP-C88F70/2020\|EPI_ISL_756495\|20 | ···················· | ···················· | ···················· | ···················· | ···················· | ···················· |
| AA | 3562 hCoV-19/England/CAMC-C860CA/2020\|EPI_ISL_756499\|20 | ···················· | ···················· | ···················· | ···················· | ···················· | ···················· |
| AA | 3563 hCoV-19/Denmark/DCGC-22617/2020\|EPI_ISL_757672\|202 | ···················· | ···················· | ···················· | ···················· | ···················· | ···················· |
| AA | 3564 hCoV-19/Denmark/DCGC-22632/2020\|EPI_ISL_757680\|202 | ···················· | ···················· | ···················· | ···················· | ···················· | ···················· |
| AA | 3565 hCoV-19/Denmark/DCGC-22634/2020\|EPI_ISL_757681\|202 | ···················· | ···················· | ···················· | ···················· | ···················· | ···················· |
| AA | 3566 hCoV-19/Denmark/DCGC-22635/2020\|EPI_ISL_757682\|202 | ···················· | ···················· | ···················· | ···················· | ···················· | ···················· |
| AA | 3567 hCoV-19/Denmark/DCGC-22639/2020\|EPI_ISL_757683\|202 | ···················· | ···················· | ···················· | ···················· | ···················· | ···················· |
| AA | 3568 hCoV-19/Denmark/DCGC-22642/2020\|EPI_ISL_757684\|202 | ···················· | ···················· | ···················· | ···················· | ···················· | ···················· |
| AA | 3569 hCoV-19/Denmark/DCGC-22644/2020\|EPI_ISL_757685\|202 | ···················· | ···················· | ···················· | ···················· | ···················· | ···················· |
| AA | 3570 hCoV-19/Denmark/DCGC-22645/2020\|EPI_ISL_757686\|202 | ···················· | ···················· | ···················· | ···················· | ···················· | ···················· |
| AA | 3571 hCoV-19/Denmark/DCGC-22648/2020\|EPI_ISL_757687\|202 | ···················· | ···················· | ···················· | ···················· | ···················· | ···················· |
| AA | 3572 hCoV-19/Denmark/DCGC-22649/2020\|EPI_ISL_757688\|202 | ···················· | ···················· | ···················· | ···················· | ···················· | ···················· |
| AA | 3573 hCoV-19/Denmark/DCGC-22650/2020\|EPI_ISL_757689\|202 | ···················· | ···················· | ···················· | ···················· | ···················· | ···················· |
| AA | 3574 hCoV-19/Denmark/DCGC-22651/2020\|EPI_ISL_757690\|202 | ···················· | ···················· | ···················· | ···················· | ···················· | ···················· |
| AA | 3575 hCoV-19/Denmark/DCGC-22655/2020\|EPI_ISL_757691\|202 | ···················· | ···················· | ···················· | ···················· | ···················· | ···················· |
| AA | 3576 hCoV-19/Denmark/DCGC-22656/2020\|EPI_ISL_757692\|202 | ···················· | ···················· | ···················· | ···················· | ···················· | ···················· |
| AA | 3577 hCoV-19/Denmark/DCGC-22657/2020\|EPI_ISL_757693\|202 | ···················· | ···················· | ···················· | ···················· | ···················· | ···················· |
| AA | 3578 hCoV-19/Denmark/DCGC-22658/2020\|EPI_ISL_757694\|202 | ···················· | ···················· | ···················· | ···················· | ···················· | ···················· |
| AA | 3579 hCoV-19/Denmark/DCGC-22659/2020\|EPI_ISL_757695\|202 | ···················· | ···················· | ···················· | ···················· | ···················· | ···················· |
| AA | 3580 hCoV-19/Denmark/DCGC-22660/2020\|EPI_ISL_757696\|202 | ···················· | ···················· | ···················· | ···················· | ···················· | ···················· |
| AA | 3581 hCoV-19/Denmark/DCGC-22661/2020\|EPI_ISL_757697\|202 | ···················· | ···················· | ···················· | ···················· | ···················· | ···················· |
| AA | 3582 hCoV-19/Denmark/DCGC-22662/2020\|EPI_ISL_757698\|202 | ···················· | ···················· | ···················· | ···················· | ···················· | ···················· |
| AA | 3583 hCoV-19/Denmark/DCGC-22663/2020\|EPI_ISL_757699\|202 | ···················· | ···················· | ···················· | ···················· | ···················· | ···················· |
| AA | 3584 hCoV-19/Denmark/DCGC-22664/2020\|EPI_ISL_757700\|202 | ···················· | ···················· | ···················· | ···················· | ···················· | ···················· |
| AA | 3585 hCoV-19/Denmark/DCGC-20158/2020\|EPI_ISL_748020\|202 | ···················· | ···················· | ···················· | ···················· | ···················· | ···················· |
| AA | 3586 hCoV-19/Denmark/DCGC-20166/2020\|EPI_ISL_748021\|202 | ···················· | ···················· | ···················· | ···················· | ···················· | ···················· |
| AA | 3587 hCoV-19/Denmark/DCGC-20167/2020\|EPI_ISL_748022\|202 | ···················· | ···················· | ···················· | ···················· | ···················· | ···················· |
| AA | 3588 hCoV-19/Denmark/DCGC-20168/2020\|EPI_ISL_748023\|202 | ···················· | ···················· | ···················· | ···················· | ···················· | ···················· |
| AA | 3589 hCoV-19/Denmark/DCGC-20169/2020\|EPI_ISL_748024\|202 | ···················· | ···················· | ···················· | ···················· | ···················· | ···················· |
| AA | 3590 hCoV-19/Denmark/DCGC-20170/2020\|EPI_ISL_748025\|202 | ···················· | ···················· | ···················· | ···················· | ···················· | ···················· |
| AA | 3591 hCoV-19/Denmark/DCGC-20171/2020\|EPI_ISL_748026\|202 | ···················· | ···················· | ···················· | ···················· | ···················· | ···················· |
| AA | 3592 hCoV-19/Denmark/DCGC-20172/2020\|EPI_ISL_748027\|202 | ···················· | ···················· | ···················· | ···················· | ···················· | ···················· |
| AA | 3593 hCoV-19/Denmark/DCGC-20175/2020\|EPI_ISL_748028\|202 | ···················· | ···················· | ···················· | ···················· | ···················· | ···················· |
| AA | 3594 hCoV-19/Denmark/DCGC-20177/2020\|EPI_ISL_748029\|202 | ···················· | ···················· | ···················· | ···················· | ···················· | ···················· |
| AA | 3595 hCoV-19/Denmark/DCGC-17783/2020\|EPI_ISL_749300\|202 | ···················· | ···················· | ···················· | ···················· | ···················· | ···················· |
| AA | 3596 hCoV-19/Denmark/DCGC-17801/2020\|EPI_ISL_749301\|202 | ···················· | ···················· | ···················· | ···················· | ···················· | ···················· |
| AA | 3597 hCoV-19/Denmark/DCGC-17803/2020\|EPI_ISL_749302\|202 | ···················· | ···················· | ···················· | ···················· | ···················· | ···················· |
| AA | 3598 hCoV-19/Denmark/DCGC-17820/2020\|EPI_ISL_749303\|202 | ···················· | ···················· | ···················· | ···················· | ···················· | ···················· |
| AA | 3599 hCoV-19/Denmark/DCGC-17843/2020\|EPI_ISL_749304\|202 | ···················· | ···················· | ···················· | ···················· | ···················· | ···················· |
| AA | 3600 hCoV-19/Denmark/DCGC-17876/2020\|EPI_ISL_749305\|202 | ···················· | ···················· | ···················· | ···················· | ···················· | ···················· |
| AA | 3601 hCoV-19/Denmark/DCGC-17917/2020\|EPI_ISL_749306\|202 | ···················· | ···················· | ···················· | ···················· | ···················· | ···················· |
| AA | 3602 hCoV-19/Denmark/DCGC-17932/2020\|EPI_ISL_749307\|202 | ···················· | ···················· | ···················· | ···················· | ···················· | ···················· |
| AA | 3603 hCoV-19/Denmark/DCGC-17959/2020\|EPI_ISL_749308\|202 | ···················· | ···················· | ···················· | ···················· | ···················· | ···················· |

AA 3604 hCoV-19/Denmark/DCGC-17996/2020|EPI_ISL_749309|202 .................... .................... ..................... .................... .................... ....................
AA 3605 hCoV-19/Denmark/DCGC-18036/2020|EPI_ISL_749310|202 .................... .................... ..................... .................... .................... ....................
AA 3606 hCoV-19/Denmark/DCGC-18087/2020|EPI_ISL_749311|202 .................... .................... ..................... .................... .................... ....................
AA 3607 hCoV-19/Denmark/DCGC-18095/2020|EPI_ISL_749312|202 .................... .................... ..................... .................... .................... ....................
AA 3608 hCoV-19/Denmark/DCGC-18119/2020|EPI_ISL_749313|202 .................... .................... ..................... .................... .................... ....................
AA 3609 hCoV-19/Denmark/DCGC-18126/2020|EPI_ISL_749314|202 .................... .................... ..................... .................... .................... ....................
AA 3610 hCoV-19/Denmark/DCGC-18131/2020|EPI_ISL_749315|202 .................... .................... ..................... .................... .................... ....................
AA 3611 hCoV-19/Denmark/DCGC-18143/2020|EPI_ISL_749316|202 .................... .................... ..................... .................... .................... ....................
AA 3612 hCoV-19/Denmark/DCGC-18149/2020|EPI_ISL_749317|202 .................... .................... ..................... .................... .................... ....................
AA 3613 hCoV-19/Denmark/DCGC-18161/2020|EPI_ISL_749318|202 .................... .................... ..................... .................... .................... ....................
AA 3614 hCoV-19/Denmark/DCGC-18230/2020|EPI_ISL_749319|202 .................... .................... ..................... .................... .................... ....................
AA 3615 hCoV-19/Denmark/DCGC-18708/2020|EPI_ISL_749350|202 .................... .................... ..................... .................... .................... ....................
AA 3616 hCoV-19/Denmark/DCGC-18730/2020|EPI_ISL_749351|202 .................... .................... ..................... .................... .................... ....................
AA 3617 hCoV-19/Denmark/DCGC-18731/2020|EPI_ISL_749352|202 .................... .................... ..................... .................... .................... ....................
AA 3618 hCoV-19/Denmark/DCGC-18740/2020|EPI_ISL_749353|202 .................... .................... ..................... .................... .................... ....................
AA 3619 hCoV-19/Denmark/DCGC-18747/2020|EPI_ISL_749354|202 .................... .................... ..................... .................... .................... ....................
AA 3620 hCoV-19/Denmark/DCGC-18034/2020|EPI_ISL_749355|202 .................... .................... ..................... .................... .................... ....................
AA 3621 hCoV-19/Denmark/DCGC-17800/2020|EPI_ISL_749356|202 .................... .................... ..................... .................... .................... ....................
AA 3622 hCoV-19/Denmark/DCGC-18702/2020|EPI_ISL_749357|202 .................... .................... ..................... .................... .................... ....................
AA 3623 hCoV-19/Denmark/DCGC-16785/2020|EPI_ISL_750301|202 .................... .................... ..................... .................... .................... ....................
AA 3624 hCoV-19/Denmark/DCGC-16814/2020|EPI_ISL_750302|202 .................... .................... ..................... .................... .................... ....................
AA 3625 hCoV-19/Denmark/DCGC-16820/2020|EPI_ISL_750303|202 .................... .................... ..................... .................... .................... ....................
AA 3626 hCoV-19/Denmark/DCGC-16845/2020|EPI_ISL_750304|202 .................... .................... ..................... .................... .................... ....................
AA 3627 hCoV-19/Denmark/DCGC-16846/2020|EPI_ISL_750305|202 .................... .................... ..................... .................... .................... ....................
AA 3628 hCoV-19/Denmark/DCGC-16858/2020|EPI_ISL_750306|202 .................... .................... ..................... .................... .................... ....................
AA 3629 hCoV-19/Denmark/DCGC-16872/2020|EPI_ISL_750307|202 .................... .................... ..................... .................... .................... ....................
AA 3630 hCoV-19/Denmark/DCGC-16873/2020|EPI_ISL_750308|202 .................... .................... ..................... .................... .................... ....................
AA 3631 hCoV-19/Denmark/DCGC-16886/2020|EPI_ISL_750309|202 .................... .................... ..................... .................... .................... ....................
AA 3632 hCoV-19/Denmark/DCGC-16908/2020|EPI_ISL_750310|202 .................... .................... ..................... .................... .................... ....................
AA 3633 hCoV-19/Denmark/DCGC-16920/2020|EPI_ISL_750311|202 .................... .................... ..................... .................... .................... ....................
AA 3634 hCoV-19/Denmark/DCGC-16930/2020|EPI_ISL_750312|202 .................... .................... ..................... .................... .................... ....................
AA 3635 hCoV-19/Denmark/DCGC-16931/2020|EPI_ISL_750313|202 .................... .................... ..................... .................... .................... ....................
AA 3636 hCoV-19/Denmark/DCGC-16934/2020|EPI_ISL_750314|202 .................... .................... ..................... .................... .................... ....................
AA 3637 hCoV-19/Denmark/DCGC-16940/2020|EPI_ISL_750315|202 .................... .................... ..................... .................... .................... ....................
AA 3638 hCoV-19/Denmark/DCGC-16943/2020|EPI_ISL_750316|202 .................... .................... ..................... .................... .................... ....................
AA 3639 hCoV-19/Denmark/DCGC-16994/2020|EPI_ISL_750317|202 .................... .................... ..................... .................... .................... ....................
AA 3640 hCoV-19/Denmark/DCGC-17006/2020|EPI_ISL_750318|202 .................... .................... ..................... .................... .................... ....................
AA 3641 hCoV-19/Denmark/DCGC-17010/2020|EPI_ISL_750319|202 .................... .................... ..................... .................... .................... ....................
AA 3642 hCoV-19/Denmark/DCGC-17022/2020|EPI_ISL_750320|202 .................... .................... ..................... .................... .................... ....................
AA 3643 hCoV-19/Denmark/DCGC-17029/2020|EPI_ISL_750321|202 .................... .................... ..................... .................... .................... ....................
AA 3644 hCoV-19/Denmark/DCGC-17031/2020|EPI_ISL_750322|202 .................... .................... ..................... .................... .................... ....................
AA 3645 hCoV-19/Denmark/DCGC-17032/2020|EPI_ISL_750323|202 .................... .................... ..................... .................... .................... ....................
AA 3646 hCoV-19/Denmark/DCGC-17038/2020|EPI_ISL_750324|202 .................... .................... ..................... .................... .................... ....................
AA 3647 hCoV-19/Denmark/DCGC-17049/2020|EPI_ISL_750325|202 .................... .................... ..................... .................... .................... ....................
AA 3648 hCoV-19/Denmark/DCGC-17086/2020|EPI_ISL_750326|202 .................... .................... ..................... .................... .................... ....................
AA 3649 hCoV-19/Denmark/DCGC-17089/2020|EPI_ISL_750327|202 .................... .................... ..................... .................... .................... ....................
AA 3650 hCoV-19/Denmark/DCGC-17109/2020|EPI_ISL_750328|202 .................... .................... ..................... .................... .................... ....................
AA 3651 hCoV-19/Denmark/DCGC-17113/2020|EPI_ISL_750329|202 .................... .................... ..................... .................... .................... ....................
AA 3652 hCoV-19/Denmark/DCGC-17125/2020|EPI_ISL_750330|202 .................... .................... ..................... .................... .................... ....................
AA 3653 hCoV-19/Denmark/DCGC-17131/2020|EPI_ISL_750331|202 .................... .................... ..................... .................... .................... ....................
AA 3654 hCoV-19/Denmark/DCGC-17142/2020|EPI_ISL_750332|202 .................... .................... ..................... .................... .................... ....................
AA 3655 hCoV-19/Denmark/DCGC-17145/2020|EPI_ISL_750333|202 .................... .................... ..................... .................... .................... ....................
AA 3656 hCoV-19/Denmark/DCGC-17163/2020|EPI_ISL_750334|202 .................... .................... ..................... .................... .................... ....................
AA 3657 hCoV-19/Denmark/DCGC-17166/2020|EPI_ISL_750335|202 .................... .................... ..................... .................... .................... ....................
AA 3658 hCoV-19/Denmark/DCGC-17172/2020|EPI_ISL_750336|202 .................... .................... ..................... .................... .................... ....................
AA 3659 hCoV-19/Denmark/DCGC-17176/2020|EPI_ISL_750337|202 .................... .................... ..................... .................... .................... ....................
AA 3660 hCoV-19/Denmark/DCGC-17188/2020|EPI_ISL_750338|202 .................... .................... ..................... .................... .................... ....................
AA 3661 hCoV-19/Denmark/DCGC-17192/2020|EPI_ISL_750339|202 .................... .................... ..................... .................... .................... ....................
AA 3662 hCoV-19/Denmark/DCGC-17197/2020|EPI_ISL_750340|202 .................... .................... ..................... .................... .................... ....................
AA 3663 hCoV-19/Denmark/DCGC-17231/2020|EPI_ISL_750341|202 .................... .................... ..................... .................... .................... ....................
AA 3664 hCoV-19/Denmark/DCGC-17240/2020|EPI_ISL_750342|202 .................... .................... ..................... .................... .................... ....................
AA 3665 hCoV-19/Denmark/DCGC-17249/2020|EPI_ISL_750343|202 .................... .................... ..................... .................... .................... ....................
AA 3666 hCoV-19/Denmark/DCGC-17260/2020|EPI_ISL_750344|202 .................... .................... ..................... .................... .................... ....................
AA 3667 hCoV-19/USA/IN-CDC-2-3714092/2020|EPI_ISL_751632|2 .................... .................... ..................... .................... .................... ....................
AA 3668 hCoV-19/USA/MT-CDC-2-3714096/2020|EPI_ISL_751633|2 .................... .................... ..................... .................... .................... ....................
AA 3669 hCoV-19/USA/MO-CDC-2-3714098/2020|EPI_ISL_751634|2 .................... .................... ..................... .................... .................... ....................
AA 3670 hCoV-19/USA/MO-CDC-2-3714099/2020|EPI_ISL_751635|2 .................... .................... ..................... .................... .................... ....................
AA 3671 hCoV-19/USA/NV-CDC-2-3714152/2020|EPI_ISL_751636|2 .................... .................... ..................... .................... .................... ....................
AA 3672 hCoV-19/USA/NV-CDC-2-3714159/2020|EPI_ISL_751637|2 .................... .A.................. ..................... .................... .................... ....................
AA 3673 hCoV-19/USA/NE-CDC-2-3714180/2020|EPI_ISL_751638|2 .................... .................... ..................... .................... .................... ....................
AA 3674 hCoV-19/USA/NV-CDC-2-3714199/2020|EPI_ISL_751639|2 .................... .A.................. ..................... .................... .................... ....................
AA 3675 hCoV-19/USA/MT-CDC-2-3714260/2020|EPI_ISL_751640|2 .................... .................... ..................... .................... .................... ....................
AA 3676 hCoV-19/USA/MN-CDC-2-3714282/2020|EPI_ISL_751641|2 .................... .................... ..................... .................... .................... ....................
AA 3677 hCoV-19/USA/NH-CDC-2-3714285/2020|EPI_ISL_751642|2 .................... .A.................. ..................... .................... .................... ....................
AA 3678 hCoV-19/USA/WY-CDC-2-3714354/2020|EPI_ISL_751643|2 .................... .................... ..................... .................... .................... ....................
AA 3679 hCoV-19/USA/TX-CDC-2-3714374/2020|EPI_ISL_751644|2 .................... .A.................. ..................... .................... .................... ....................
AA 3680 hCoV-19/USA/TX-CDC-2-3714383/2020|EPI_ISL_751645|2 .................... .................... ..................... .................... .................... ....................
AA 3681 hCoV-19/USA/TX-CDC-2-3714392/2020|EPI_ISL_751646|2 .................... .................... ..................... .................... .................... ....................
AA 3682 hCoV-19/USA/AL-CDC-2-3714561/2020|EPI_ISL_751647|2 .................... .................... ..................... .................... .................... ....................
AA 3683 hCoV-19/USA/KY-CDC-2-3714571/2020|EPI_ISL_751648|2 .................... .................... ..................... .................... .................... ....................

| | | | | | | | |
|---|---|---|---|---|---|---|---|
| AA | 3684 hCoV-19/USA/CT-CDC-2-3713968/2020\|EPI_ISL_751649\|2 | ···················· | ···················· | ···················· | ···················· | ···················· | ···················· |
| AA | 3685 hCoV-19/USA/CT-CDC-2-3713977/2020\|EPI_ISL_751650\|2 | ···················· | ···················· | ···················· | ···················· | ···················· | ···················· |
| AA | 3686 hCoV-19/USA/HI-CDC-2-3713978/2020\|EPI_ISL_751651\|2 | ···················· | ·A·················· | ···················· | ···················· | ···················· | ···················· |
| AA | 3687 hCoV-19/USA/AR-CDC-2-3713986/2020\|EPI_ISL_751652\|2 | ···················· | ···················· | ···················· | ···················· | ···················· | ···················· |
| AA | 3688 hCoV-19/USA/MN-CDC-2-3713988/2020\|EPI_ISL_751653\|2 | ···················· | ···················· | ···················· | ···················· | ···················· | ···················· |
| AA | 3689 hCoV-19/USA/MN-CDC-2-3714089/2020\|EPI_ISL_751654\|2 | ···················· | ···················· | ···················· | ···················· | ···················· | ···················· |
| AA | 3690 hCoV-19/USA/MN-CDC-2-3714097/2020\|EPI_ISL_751655\|2 | ···················· | ···················· | ···················· | ···················· | ···················· | ···················· |
| AA | 3691 hCoV-19/USA/MO-CDC-2-3714153/2020\|EPI_ISL_751656\|2 | ···················· | ···················· | ···················· | ···················· | ···················· | ···················· |
| AA | 3692 hCoV-19/USA/MO-CDC-2-3714154/2020\|EPI_ISL_751657\|2 | ···················· | ···················· | ···················· | ···················· | ···················· | ···················· |
| AA | 3693 hCoV-19/USA/NY-CDC-2-3714160/2020\|EPI_ISL_751658\|2 | ···················· | ···················· | ···················· | ···················· | ···················· | ···················· |
| AA | 3694 hCoV-19/USA/NJ-CDC-2-3714169/2020\|EPI_ISL_751659\|2 | ···················· | ···················· | ···················· | ···················· | ···················· | ···················· |
| AA | 3695 hCoV-19/USA/MN-CDC-2-3714175/2020\|EPI_ISL_751660\|2 | ···················· | ·A·················· | ···················· | ···················· | ···················· | ···················· |
| AA | 3696 hCoV-19/USA/MS-CDC-2-3714183/2020\|EPI_ISL_751661\|2 | ···················· | ···················· | ···················· | ···················· | ···················· | ···················· |
| AA | 3697 hCoV-19/USA/NJ-CDC-2-3714252/2020\|EPI_ISL_751662\|2 | ···················· | ·A·················· | ···················· | ···················· | ···················· | ···················· |
| AA | 3698 hCoV-19/USA/WY-CDC-2-3714296/2020\|EPI_ISL_751663\|2 | ···················· | ·A·················· | ···················· | ···················· | ···················· | ···················· |
| AA | 3699 hCoV-19/USA/SC-CDC-2-3714297/2020\|EPI_ISL_751664\|2 | ···················· | ···················· | ···················· | ···················· | ···················· | ···················· |
| AA | 3700 hCoV-19/USA/SC-CDC-2-3714365/2020\|EPI_ISL_751665\|2 | ···················· | ·A·················· | ···················· | ···················· | ···················· | ···················· |
| AA | 3701 hCoV-19/USA/IA-CDC-2-3714396/2020\|EPI_ISL_751666\|2 | ···················· | ···················· | ···················· | ···················· | ···················· | ···················· |
| AA | 3702 hCoV-19/USA/RI-CDC-2-3714491/2020\|EPI_ISL_751667\|2 | ···················· | ···················· | ···················· | ···················· | ···················· | ···················· |
| AA | 3703 hCoV-19/USA/AL-CDC-2-3714551/2020\|EPI_ISL_751668\|2 | ···················· | ···················· | ···················· | ···················· | ···················· | ···················· |
| AA | 3704 hCoV-19/USA/LA-CDC-2-3714562/2020\|EPI_ISL_751669\|2 | ········T··········· | ·A·················· | ···················· | ···················· | ···················· | ···················· |
| AA | 3705 hCoV-19/USA/KY-CDC-2-3714580/2020\|EPI_ISL_751670\|2 | ···················· | ···················· | ···················· | ···················· | ···················· | ···················· |
| AA | 3706 hCoV-19/USA/RI-CDC-2-3714588/2020\|EPI_ISL_751671\|2 | ···················· | ···················· | ···················· | ···················· | ···················· | ···················· |
| AA | 3707 hCoV-19/USA/IN-CDC-2-3713970/2020\|EPI_ISL_751672\|2 | ···················· | ···················· | ···················· | ···················· | ···················· | ···················· |
| AA | 3708 hCoV-19/USA/HI-CDC-2-3713981/2020\|EPI_ISL_751673\|2 | ···················· | ·A·················· | ···················· | ···················· | ···················· | ···················· |
| AA | 3709 hCoV-19/USA/HI-CDC-2-3713991/2020\|EPI_ISL_751674\|2 | ···················· | ·A·················· | ···················· | ···················· | ···················· | ···················· |
| AA | 3710 hCoV-19/USA/HI-CDC-2-3713994/2020\|EPI_ISL_751675\|2 | ···················· | ·A·················· | ···················· | ···················· | ···················· | ···················· |
| AA | 3711 hCoV-19/USA/HI-H200344/2020\|EPI_ISL_752900\|2020-08 | ···················· | ·A·················· | ···················· | ···················· | ···················· | ···················· |
| AA | 3712 hCoV-19/USA/HI-H200345/2020\|EPI_ISL_752901\|2020-08 | ···················· | ·A·················· | ···················· | ···················· | ···················· | ···················· |
| AA | 3713 hCoV-19/USA/HI-H200346/2020\|EPI_ISL_752902\|2020-09 | ···················· | ·A·················· | ···················· | ···················· | ···················· | ···················· |
| AA | 3714 hCoV-19/USA/HI-H200347/2020\|EPI_ISL_752903\|2020-09 | ···················· | ·A·················· | ···················· | ···················· | ···················· | ···················· |
| AA | 3715 hCoV-19/USA/HI-H200348/2020\|EPI_ISL_752904\|2020-09 | ···················· | ·A·················· | ···················· | ···················· | ···················· | ···················· |
| AA | 3716 hCoV-19/USA/HI-H200349/2020\|EPI_ISL_752905\|2020-09 | ···················· | ·A·················· | ···················· | ···················· | ···················· | ···················· |
| AA | 3717 hCoV-19/USA/HI-H200350/2020\|EPI_ISL_752906\|2020-09 | ···················· | ·A·················· | ···················· | ···················· | ···················· | ···················· |
| AA | 3718 hCoV-19/USA/HI-H200351/2020\|EPI_ISL_752907\|2020-09 | ···················· | ·A·················· | ···················· | ···················· | ···················· | ···················· |
| AA | 3719 hCoV-19/USA/HI-H200353/2020\|EPI_ISL_752909\|2020-09 | ···················· | ·A·················· | ···················· | ···················· | ···················· | ···················· |
| AA | 3720 hCoV-19/USA/HI-H200354/2020\|EPI_ISL_752910\|2020-09 | ···················· | ·A·················· | ···················· | ···················· | ···················· | ···················· |
| AA | 3721 hCoV-19/USA/HI-H200355/2020\|EPI_ISL_752911\|2020-09 | ···················· | ·A·················· | ···················· | ···················· | ···················· | ···················· |
| AA | 3722 hCoV-19/USA/HI-H200356/2020\|EPI_ISL_752912\|2020-09 | ···················· | ·A·················· | ···················· | ···················· | ···················· | ···················· |
| AA | 3723 hCoV-19/USA/HI-H200357/2020\|EPI_ISL_752913\|2020-09 | ···················· | ·A·················· | ···················· | ···················· | ···················· | ···················· |
| AA | 3724 hCoV-19/USA/HI-H200358/2020\|EPI_ISL_752914\|2020-09 | ···················· | ·A·················· | ···················· | ···················· | ···················· | ···················· |
| AA | 3725 hCoV-19/USA/HI-H200359/2020\|EPI_ISL_752915\|2020-09 | ···················· | ·A·················· | ···················· | ···················· | ···················· | ···················· |
| AA | 3726 hCoV-19/USA/HI-H200360/2020\|EPI_ISL_752916\|2020-09 | ···················· | ·A·················· | ···················· | ···················· | ···················· | ···················· |
| AA | 3727 hCoV-19/USA/HI-H200361/2020\|EPI_ISL_752917\|2020-09 | ···················· | ·A·················· | ···················· | ···················· | ···················· | ···················· |
| AA | 3728 hCoV-19/USA/HI-H200362/2020\|EPI_ISL_752918\|2020-09 | ···················· | ·A·················· | ···················· | ···················· | ···················· | ···················· |
| AA | 3729 hCoV-19/USA/HI-H200363/2020\|EPI_ISL_752919\|2020-09 | ···················· | ·A·················· | ···················· | ···················· | ···················· | ···················· |
| AA | 3730 hCoV-19/USA/HI-H200364/2020\|EPI_ISL_752920\|2020-09 | ···················· | ·A·················· | ···················· | ···················· | ···················· | ···················· |
| AA | 3731 hCoV-19/USA/HI-H200365/2020\|EPI_ISL_752921\|2020-09 | ···················· | ·A·················· | ···················· | ···················· | ···················· | ···················· |
| AA | 3732 hCoV-19/USA/HI-H200366/2020\|EPI_ISL_752922\|2020-09 | ···················· | ·A·················· | ···················· | ···················· | ···················· | ···················· |
| AA | 3733 hCoV-19/USA/HI-H200367/2020\|EPI_ISL_752923\|2020-09 | ···················· | ·A·················· | ···················· | ···················· | ···················· | ···················· |
| AA | 3734 hCoV-19/USA/HI-H200368/2020\|EPI_ISL_752924\|2020-09 | ···················· | ·A·················· | ···················· | ···················· | ···················· | ···················· |
| AA | 3735 hCoV-19/USA/HI-H200376/2020\|EPI_ISL_752925\|2020-09 | ···················· | ·A·················· | ···················· | ···················· | ···················· | ···················· |
| AA | 3736 hCoV-19/USA/HI-H200377/2020\|EPI_ISL_752926\|2020-09 | ···················· | ·A·················· | ···················· | ···················· | ···················· | ···················· |
| AA | 3737 hCoV-19/USA/HI-H200379/2020\|EPI_ISL_752927\|2020-09 | ···················· | ·A·················· | ···················· | ···················· | ···················· | ···················· |
| AA | 3738 hCoV-19/USA/HI-H200381/2020\|EPI_ISL_752928\|2020-09 | ···················· | ·A·················· | ···················· | ···················· | ···················· | ···················· |
| AA | 3739 hCoV-19/USA/HI-H200382/2020\|EPI_ISL_752929\|2020-09 | ···················· | ·A·················· | ···················· | ···················· | ···················· | ···················· |
| AA | 3740 hCoV-19/USA/HI-H200419/2020\|EPI_ISL_752963\|2020-08 | ···················· | ·A·················· | ···················· | ···················· | ···················· | ···················· |
| AA | 3741 hCoV-19/USA/HI-H200420/2020\|EPI_ISL_752964\|2020-08 | ···················· | ·A·················· | ···················· | ···················· | ···················· | ···················· |
| AA | 3742 hCoV-19/USA/HI-H200421/2020\|EPI_ISL_752965\|2020-08 | ···················· | ·A·················· | ···················· | ···················· | ···················· | ···················· |
| AA | 3743 hCoV-19/USA/HI-H200422/2020\|EPI_ISL_752966\|2020-08 | ···················· | ·A·················· | ···················· | ···················· | ···················· | ···················· |
| AA | 3744 hCoV-19/USA/HI-H200423/2020\|EPI_ISL_752967\|2020-09 | ···················· | ·A·················· | ···················· | ···················· | ···················· | ···················· |
| AA | 3745 hCoV-19/USA/HI-H200424/2020\|EPI_ISL_752968\|2020-09 | ···················· | ·A·················· | ···················· | ···················· | ···················· | ···················· |
| AA | 3746 hCoV-19/USA/HI-H200425/2020\|EPI_ISL_752969\|2020-09 | ···················· | ·A·················· | ···················· | ···················· | ···················· | ···················· |
| AA | 3747 hCoV-19/USA/HI-H200426/2020\|EPI_ISL_752970\|2020-09 | ···················· | ·A·················· | ···················· | ···················· | ···················· | ···················· |
| AA | 3748 hCoV-19/USA/HI-H200427/2020\|EPI_ISL_752971\|2020-09 | ···················· | ·A·················· | ···················· | ···················· | ···················· | ···················· |
| AA | 3749 hCoV-19/USA/HI-H200428/2020\|EPI_ISL_752972\|2020-09 | ···················· | ·A·················· | ···················· | ···················· | ···················· | ···················· |
| AA | 3750 hCoV-19/USA/HI-H200430/2020\|EPI_ISL_752974\|2020-09 | ···················· | ·A·················· | ···················· | ···················· | ···················· | ···················· |
| AA | 3751 hCoV-19/USA/HI-H200431/2020\|EPI_ISL_752975\|2020-10 | ···················· | ·A·················· | ···················· | ···················· | ···················· | ···················· |
| AA | 3752 hCoV-19/USA/HI-H200433/2020\|EPI_ISL_752976\|2020-10 | ···················· | ·A·················· | ···················· | ···················· | ···················· | ···················· |
| AA | 3753 hCoV-19/USA/HI-H200434/2020\|EPI_ISL_752977\|2020-10 | ···················· | ·A·················· | ···················· | ···················· | ···················· | ···················· |
| AA | 3754 hCoV-19/USA/HI-H200435/2020\|EPI_ISL_752978\|2020-10 | ···················· | ·A·················· | ···················· | ···················· | ···················· | ···················· |
| AA | 3755 hCoV-19/USA/HI-H200436/2020\|EPI_ISL_752979\|2020-10 | ···················· | ·A·················· | ···················· | ···················· | ···················· | ···················· |
| AA | 3756 hCoV-19/USA/HI-H200437/2020\|EPI_ISL_752980\|2020-10 | ···················· | ·A·················· | ···················· | ···················· | ···················· | ···················· |
| AA | 3757 hCoV-19/USA/HI-H200438/2020\|EPI_ISL_752981\|2020-10 | ···················· | ·A·················· | ···················· | ···················· | ···················· | ···················· |
| AA | 3758 hCoV-19/USA/HI-H200439/2020\|EPI_ISL_752982\|2020-10 | ···················· | ·A·················· | ···················· | ···················· | ···················· | ···················· |
| AA | 3759 hCoV-19/USA/HI-H200440/2020\|EPI_ISL_752983\|2020-10 | ···················· | ·A·················· | ···················· | ···················· | ···················· | ···················· |
| AA | 3760 hCoV-19/USA/HI-H200441/2020\|EPI_ISL_752984\|2020-10 | ···················· | ·A·················· | ···················· | ···················· | ···················· | ···················· |
| AA | 3761 hCoV-19/USA/HI-H200442/2020\|EPI_ISL_752985\|2020-10 | ···················· | ·A·················· | ···················· | ···················· | ···················· | ···················· |
| AA | 3762 hCoV-19/USA/HI-H200443/2020\|EPI_ISL_752986\|2020-10 | ···················· | ·A·················· | ···················· | ···················· | ···················· | ···················· |
| AA | 3763 hCoV-19/USA/HI-H200444/2020\|EPI_ISL_752987\|2020-10 | ···················· | ·A·················· | ···················· | ···················· | ···················· | ···················· |

AA 3764 hCoV-19/USA/HI-H200445/2020|EPI_ISL_752988|2020-10 ···················· ·A·················· ····················· ···················· ···················· ····················
AA 3765 hCoV-19/USA/HI-H200446/2020|EPI_ISL_752989|2020-10 ···················· ·A·················· ····················· ···················· ···················· ····················
AA 3766 hCoV-19/USA/HI-H200447/2020|EPI_ISL_752990|2020-10 ···················· ·A·················· ····················· ···················· ···················· ····················
AA 3767 hCoV-19/USA/HI-H200448/2020|EPI_ISL_752991|2020-10 ···················· ·A·················· ····················· ···················· ···················· ····················
AA 3768 hCoV-19/USA/HI-H200449/2020|EPI_ISL_752992|2020-10 ···················· ·A·················· ····················· ···················· ···················· ····················
AA 3769 hCoV-19/USA/HI-H200450/2020|EPI_ISL_752993|2020-10 ···················· ·A·················· ····················· ···················· ···················· ····················
AA 3770 hCoV-19/USA/HI-H200451/2020|EPI_ISL_752994|2020-10 ···················· ·A·················· ····················· ···················· ···················· ····················
AA 3771 hCoV-19/USA/HI-H200452/2020|EPI_ISL_752995|2020-10 ···················· ·A·················· ····················· ···················· ···················· ····················
AA 3772 hCoV-19/USA/HI-H200453/2020|EPI_ISL_752996|2020-09 ···················· ·A·················· ····················· ···················· ···················· ····················
AA 3773 hCoV-19/USA/HI-H200454/2020|EPI_ISL_752997|2020-09 ···················· ·A·················· ····················· ···················· ···················· ····················
AA 3774 hCoV-19/USA/HI-H200455/2020|EPI_ISL_752998|2020-09 ···················· ·A·················· ····················· ···················· ···················· ····················
AA 3775 hCoV-19/USA/HI-H200456/2020|EPI_ISL_752999|2020-09 ···················· ·A·················· ····················· ···················· ···················· ····················
AA 3776 hCoV-19/USA/CA-SEARCH-5516/2020|EPI_ISL_755136|202 ···················· ···················· ····················· ···················· ···················· ····················
AA 3777 hCoV-19/USA/CA-SEARCH-5385/2020|EPI_ISL_755137|202 ···················· ···················· ····················· ···················· ···················· ····················
AA 3778 hCoV-19/USA/CA-SEARCH-5505/2020|EPI_ISL_755138|202 ···················· ·A·················· ····················· ···················· ···················· ····················
AA 3779 hCoV-19/USA/CA-SEARCH-5509/2020|EPI_ISL_755139|202 ···················· ·A·················· ····················· ···················· ···················· ····················
AA 3780 hCoV-19/USA/CA-SEARCH-5336/2020|EPI_ISL_755140|202 ···················· ·A·················· ····················· ···················· ···················· ····················
AA 3781 hCoV-19/Jordan/SEARCH-5433/2020|EPI_ISL_755141|202 ···················· ···················· ····················· ···················· ···················· ····················
AA 3782 hCoV-19/Jordan/SEARCH-5464/2020|EPI_ISL_755142|202 ···················· ···················· ····················· ···················· ···················· ····················
AA 3783 hCoV-19/Jordan/SEARCH-5469/2020|EPI_ISL_755143|202 ···················· ···················· ····················· ···················· ···················· ····················
AA 3784 hCoV-19/Jordan/SEARCH-5470/2020|EPI_ISL_755144|202 ···················· ···················· ····················· ···················· ······T············· ····················
AA 3785 hCoV-19/Jordan/SEARCH-5487/2020|EPI_ISL_755145|202 ···················· ···················· ····················· ···················· ···················· ····················
AA 3786 hCoV-19/USA/CA-SEARCH-5328/2020|EPI_ISL_755147|202 ···················· ·A·················· ····················· ···················· ···················· ····················
AA 3787 hCoV-19/USA/CA-SEARCH-5329/2020|EPI_ISL_755148|202 ···················· ·A·················· ····················· ···················· ···················· ····················
AA 3788 hCoV-19/USA/CA-SEARCH-5330/2020|EPI_ISL_755149|202 ···················· ···················· ····················· ···················· ···················· ····················
AA 3789 hCoV-19/USA/CA-SEARCH-5331/2020|EPI_ISL_755150|202 ···················· ·A·················· ····················· ···················· ···················· ····················
AA 3790 hCoV-19/USA/CA-SEARCH-5332/2020|EPI_ISL_755151|202 ···················· ·A·················· ····················· ···················· ···················· ····················
AA 3791 hCoV-19/USA/CA-SEARCH-5333/2020|EPI_ISL_755152|202 ···················· ·A·················· ····················· ···················· ···················· ····················
AA 3792 hCoV-19/USA/CA-SEARCH-5334/2020|EPI_ISL_755153|202 ···················· ·A·················· ····················· ···················· ···················· ····················
AA 3793 hCoV-19/USA/CA-SEARCH-5335/2020|EPI_ISL_755154|202 ···················· ···················· ····················· ···················· ···················· ····················
AA 3794 hCoV-19/USA/CA-SEARCH-5337/2020|EPI_ISL_755155|202 ···················· ···················· ····················· ···················· ···················· ····················
AA 3795 hCoV-19/USA/CA-SEARCH-5339/2020|EPI_ISL_755156|202 ···················· ·A·················· ····················· ···················· ···················· ····················
AA 3796 hCoV-19/USA/CA-SEARCH-5340/2020|EPI_ISL_755157|202 ···················· ···················· ····················· ···················· ···················· ····················
AA 3797 hCoV-19/USA/CA-SEARCH-5341/2020|EPI_ISL_755158|202 ···················· ···················· ····················· ···················· ···················· ····················
AA 3798 hCoV-19/USA/CA-SEARCH-5342/2020|EPI_ISL_755159|202 ···················· ·A·················· ····················· ···················· ···················· ····················
AA 3799 hCoV-19/USA/CA-SEARCH-5359/2020|EPI_ISL_755171|202 ···················· ·A·················· ····················· ···················· ···················· ····················
AA 3800 hCoV-19/USA/CA-SEARCH-5360/2020|EPI_ISL_755172|202 ···················· ·A·················· ····················· ···················· ···················· ····················
AA 3801 hCoV-19/USA/CA-SEARCH-5361/2020|EPI_ISL_755173|202 ············T······· ···················· ····················· ···················· ···················· ····················
AA 3802 hCoV-19/USA/CA-SEARCH-5362/2020|EPI_ISL_755174|202 ···················· ···················· ····················· ···················· ···················· ····················
AA 3803 hCoV-19/USA/CA-SEARCH-5363/2020|EPI_ISL_755175|202 ···················· ···················· ····················· ···················· ···················· ····················
AA 3804 hCoV-19/USA/CA-SEARCH-5364/2020|EPI_ISL_755176|202 ···················· ···················· ····················· ···················· ···················· ····················
AA 3805 hCoV-19/USA/CA-SEARCH-5368/2020|EPI_ISL_755180|202 ···················· ···················· ····················· ···················· ···················· ····················
AA 3806 hCoV-19/USA/CA-SEARCH-5369/2020|EPI_ISL_755181|202 ···················· ···················· ····················· ···················· ···················· ····················
AA 3807 hCoV-19/USA/CA-SEARCH-5370/2020|EPI_ISL_755182|202 ···················· ···················· ····················· ···················· ···················· ····················
AA 3808 hCoV-19/USA/CA-SEARCH-5371/2020|EPI_ISL_755183|202 ···················· ···················· ····················· ···················· ···················· ····················
AA 3809 hCoV-19/USA/CA-SEARCH-5372/2020|EPI_ISL_755184|202 ···················· ···················· ····················· ···················· ···················· ····················
AA 3810 hCoV-19/USA/CA-SEARCH-5373/2020|EPI_ISL_755185|202 ···················· ·A·················· ····················· ···················· ···················· ····················
AA 3811 hCoV-19/USA/CA-SEARCH-5374/2020|EPI_ISL_755186|202 ···················· ·A·················· ····················· ···················· ···················· ····················
AA 3812 hCoV-19/USA/CA-SEARCH-5375/2020|EPI_ISL_755187|202 ···················· ···················· ····················· ···················· ···················· ····················
AA 3813 hCoV-19/USA/CA-SEARCH-5376/2020|EPI_ISL_755188|202 ···················· ···················· ····················· ···················· ···················· ····················
AA 3814 hCoV-19/USA/CA-SEARCH-5377/2020|EPI_ISL_755189|202 ···················· ···················· ····················· ···················· ···················· ····················
AA 3815 hCoV-19/USA/CA-SEARCH-5378/2020|EPI_ISL_755190|202 ···················· ···················· ····················· ···················· ···················· ····················
AA 3816 hCoV-19/USA/CA-SEARCH-5379/2020|EPI_ISL_755191|202 ···················· ·A·················· ····················· ···················· ···················· ····················
AA 3817 hCoV-19/USA/CA-SEARCH-5380/2020|EPI_ISL_755192|202 ···················· ···················· ····················· ···················· ···················· ····················
AA 3818 hCoV-19/USA/CA-SEARCH-5381/2020|EPI_ISL_755193|202 ···················· ·A·················· ····················· ···················· ···················· ····················
AA 3819 hCoV-19/USA/CA-SEARCH-5382/2020|EPI_ISL_755194|202 ···················· ···················· ····················· ···················· ···················· ····················
AA 3820 hCoV-19/USA/CA-SEARCH-5383/2020|EPI_ISL_755195|202 ···················· ···················· ····················· ···················· ···················· ····················
AA 3821 hCoV-19/USA/CA-SEARCH-5384/2020|EPI_ISL_755196|202 ···················· ···················· ····················· ···················· ···················· ····················
AA 3822 hCoV-19/USA/CA-SEARCH-5386/2020|EPI_ISL_755197|202 ···················· ·A·················· ····················· ···················· ···················· ····················
AA 3823 hCoV-19/USA/CA-SEARCH-5387/2020|EPI_ISL_755198|202 ···················· ···················· ····················· ···················· ···················· ····················
AA 3824 hCoV-19/USA/CA-SEARCH-5388/2020|EPI_ISL_755199|202 ···················· ···················· ····················· ···················· ···················· ····················
AA 3825 hCoV-19/England/ALDP-C8B6F0/2020|EPI_ISL_756450|20 ···················· ···················· ····················· ···················· ···················· ····················
AA 3826 hCoV-19/England/ALDP-C8B98E/2020|EPI_ISL_756452|20 ···················· ···················· ····················· ···················· ···················· ····················
AA 3827 hCoV-19/England/ALDP-C895C8/2020|EPI_ISL_756457|20 ···················· ···················· ····················· ···················· ···················· ····················
AA 3828 hCoV-19/England/ALDP-C8B845/2020|EPI_ISL_756458|20 ···················· ···················· ····················· ···················· ···················· ····················
AA 3829 hCoV-19/England/ALDP-C8B195/2020|EPI_ISL_756459|20 ···················· ···················· ····················· ···················· ···················· ····················
AA 3830 hCoV-19/England/ALDP-C8B687/2020|EPI_ISL_756461|20 ··G················· ···················· ····················· ···················· ···················· ····················
AA 3831 hCoV-19/England/ALDP-C8B6C3/2020|EPI_ISL_756462|20 ···················· ···················· ····················· ···················· ···················· ····················
AA 3832 hCoV-19/England/ALDP-C8959B/2020|EPI_ISL_756465|20 ···················· ·A·················· ····················· ···················· ···················· ··A·················
AA 3833 hCoV-19/England/CAMC-C84008/2020|EPI_ISL_756467|20 ···················· ···················· ····················· ···················· ···················· ····················
AA 3834 hCoV-19/England/ALDP-C88444/2020|EPI_ISL_756470|20 ···················· ···················· ····················· ···················· ···················· ····················
AA 3835 hCoV-19/England/ALDP-C88F52/2020|EPI_ISL_756471|20 ···················· ···················· ····················· ···················· ···················· ····················
AA 3836 hCoV-19/England/ALDP-C8B55D/2020|EPI_ISL_756479|20 ···················· ···················· ····················· ···················· ···················· ····················
AA 3837 hCoV-19/England/CAMC-C85007/2020|EPI_ISL_756480|20 ···················· ···················· ····················· ···················· ······T············· ····················
AA 3838 hCoV-19/England/CAMC-C866B9/2020|EPI_ISL_756481|20 ···················· ···················· ····················· ···················· ···················· ····················
AA 3839 hCoV-19/England/ALDP-C8B63C/2020|EPI_ISL_756482|20 ···················· ···················· ····················· ···················· ···················· ····················
AA 3840 hCoV-19/England/ALDP-C8B1C2/2020|EPI_ISL_756483|20 ···················· ···················· ····················· ···················· ···················· ····················
AA 3841 hCoV-19/Denmark/DCGC-20128/2020|EPI_ISL_748000|202 ···················· ···················· ····················· ···················· ···················· ····················
AA 3842 hCoV-19/Denmark/DCGC-20129/2020|EPI_ISL_748001|202 ···················· ···················· ····················· ···················· ···················· ····················
AA 3843 hCoV-19/Denmark/DCGC-20131/2020|EPI_ISL_748002|202 ···················· ···················· ····················· ···················· ···················· ····················

| | | | | | | | | |
|---|---|---|---|---|---|---|---|---|
| AA | 3844 | hCoV-19/Denmark/DCGC-20132/2020\|EPI_ISL_748003\|202 | .................... | .................... | ..................... | .................... | .................... | .................... |
| AA | 3845 | hCoV-19/Denmark/DCGC-20133/2020\|EPI_ISL_748004\|202 | .................... | .................... | ..................... | .................... | .................... | .................... |
| AA | 3846 | hCoV-19/Denmark/DCGC-20135/2020\|EPI_ISL_748005\|202 | .................... | .................... | ..................... | .................... | .................... | .................... |
| AA | 3847 | hCoV-19/Denmark/DCGC-20136/2020\|EPI_ISL_748006\|202 | .................... | .................... | ..................... | .................... | .................... | .................... |
| AA | 3848 | hCoV-19/Denmark/DCGC-20137/2020\|EPI_ISL_748007\|202 | .................... | .................... | ..................... | .................... | .................... | .................... |
| AA | 3849 | hCoV-19/Denmark/DCGC-20138/2020\|EPI_ISL_748008\|202 | .................... | .................... | ..................... | .................... | .................... | .................... |
| AA | 3850 | hCoV-19/Denmark/DCGC-20141/2020\|EPI_ISL_748009\|202 | .................... | .................... | ..................... | .................... | .................... | .................... |
| AA | 3851 | hCoV-19/Denmark/DCGC-20143/2020\|EPI_ISL_748010\|202 | .................... | .A.................. | ..................... | .................... | .................... | ..A................. |
| AA | 3852 | hCoV-19/Denmark/DCGC-20144/2020\|EPI_ISL_748011\|202 | .................... | .................... | ..................... | .................... | .................... | .................... |
| AA | 3853 | hCoV-19/Denmark/DCGC-20146/2020\|EPI_ISL_748012\|202 | .................... | .................... | ..................... | .................... | .................... | .................... |
| AA | 3854 | hCoV-19/Denmark/DCGC-20147/2020\|EPI_ISL_748013\|202 | .................... | .................... | ..................... | .................... | .................... | .................... |
| AA | 3855 | hCoV-19/Denmark/DCGC-20148/2020\|EPI_ISL_748014\|202 | .................... | .................... | ..................... | .................... | .................... | .................... |
| AA | 3856 | hCoV-19/Denmark/DCGC-20149/2020\|EPI_ISL_748015\|202 | .................... | .................... | ..................... | .................... | .................... | .................... |
| AA | 3857 | hCoV-19/Denmark/DCGC-20151/2020\|EPI_ISL_748016\|202 | .................... | .A.................. | ..................... | .................... | .................... | ..A................. |
| AA | 3858 | hCoV-19/Denmark/DCGC-20152/2020\|EPI_ISL_748017\|202 | .................... | .................... | ..................... | .................... | .................... | .................... |
| AA | 3859 | hCoV-19/Denmark/DCGC-20155/2020\|EPI_ISL_748018\|202 | .................... | .................... | ..................... | .................... | .................... | .................... |
| AA | 3860 | hCoV-19/Denmark/DCGC-20157/2020\|EPI_ISL_748019\|202 | .................... | .................... | ..................... | .................... | .................... | .................... |
| AA | 3861 | hCoV-19/Denmark/DCGC-20178/2020\|EPI_ISL_748030\|202 | .................... | .................... | ..................... | .................... | .................... | .................... |
| AA | 3862 | hCoV-19/Denmark/DCGC-20179/2020\|EPI_ISL_748031\|202 | .................... | .................... | ..................... | .................... | .................... | .................... |
| AA | 3863 | hCoV-19/Denmark/DCGC-20180/2020\|EPI_ISL_748032\|202 | .................... | .................... | ..................... | .................... | .................... | .................... |
| AA | 3864 | hCoV-19/Denmark/DCGC-20181/2020\|EPI_ISL_748033\|202 | .................... | .................... | ..................... | .................... | .................... | .................... |
| AA | 3865 | hCoV-19/Denmark/DCGC-20182/2020\|EPI_ISL_748034\|202 | .................... | .................... | ..................... | .................... | .................... | .................... |
| AA | 3866 | hCoV-19/Denmark/DCGC-20183/2020\|EPI_ISL_748035\|202 | .................... | .................... | ..................... | .................... | .................... | .................... |
| AA | 3867 | hCoV-19/Denmark/DCGC-20184/2020\|EPI_ISL_748036\|202 | .................... | .................... | ..................... | .................... | .................... | .................... |
| AA | 3868 | hCoV-19/Denmark/DCGC-20186/2020\|EPI_ISL_748037\|202 | .................... | .................... | ..................... | .................... | .................... | .................... |
| AA | 3869 | hCoV-19/Denmark/DCGC-20188/2020\|EPI_ISL_748038\|202 | .................... | .................... | ..................... | .................... | .................... | .................... |
| AA | 3870 | hCoV-19/Denmark/DCGC-20191/2020\|EPI_ISL_748039\|202 | .................... | .................... | ..................... | .................... | .................... | .................... |
| AA | 3871 | hCoV-19/Denmark/DCGC-20192/2020\|EPI_ISL_748040\|202 | .................... | .................... | ..................... | .................... | .................... | .................... |
| AA | 3872 | hCoV-19/Denmark/DCGC-20193/2020\|EPI_ISL_748041\|202 | .................... | .................... | ..................... | .................... | .................... | .................... |
| AA | 3873 | hCoV-19/Denmark/DCGC-20194/2020\|EPI_ISL_748042\|202 | .................... | .................... | ..................... | .................... | .................... | .................... |
| AA | 3874 | hCoV-19/Denmark/DCGC-20196/2020\|EPI_ISL_748043\|202 | .................... | .................... | ..................... | .................... | .................... | .................... |
| AA | 3875 | hCoV-19/Denmark/DCGC-20197/2020\|EPI_ISL_748044\|202 | .................... | .................... | ..................... | .................... | .................... | .................... |
| AA | 3876 | hCoV-19/Denmark/DCGC-20199/2020\|EPI_ISL_748045\|202 | .................... | .................... | ..................... | .................... | .................... | .................... |
| AA | 3877 | hCoV-19/Denmark/DCGC-20200/2020\|EPI_ISL_748046\|202 | .................... | .................... | ..................... | .................... | .................... | .................... |
| AA | 3878 | hCoV-19/Denmark/DCGC-20201/2020\|EPI_ISL_748047\|202 | .................... | .................... | ..................... | .................... | .................... | .................... |
| AA | 3879 | hCoV-19/Denmark/DCGC-20203/2020\|EPI_ISL_748048\|202 | .................... | .................... | ..................... | .................... | .................... | .................... |
| AA | 3880 | hCoV-19/Denmark/DCGC-20204/2020\|EPI_ISL_748049\|202 | .................... | .................... | ..................... | .................... | .................... | .................... |
| AA | 3881 | hCoV-19/Denmark/DCGC-20205/2020\|EPI_ISL_748050\|202 | .................... | .................... | ..................... | .................... | .................... | .................... |
| AA | 3882 | hCoV-19/Denmark/DCGC-20210/2020\|EPI_ISL_748051\|202 | .................... | .................... | ..................... | .................... | .................... | .................... |
| AA | 3883 | hCoV-19/Denmark/DCGC-20212/2020\|EPI_ISL_748052\|202 | .................... | .................... | ..................... | .................... | .................... | .................... |
| AA | 3884 | hCoV-19/Denmark/DCGC-20215/2020\|EPI_ISL_748053\|202 | .................... | .................... | ..................... | .................... | .................... | .................... |
| AA | 3885 | hCoV-19/Denmark/DCGC-20216/2020\|EPI_ISL_748054\|202 | .................... | .A.................. | ..................... | .................... | .................... | .................... |
| AA | 3886 | hCoV-19/Denmark/DCGC-20217/2020\|EPI_ISL_748055\|202 | .................... | .................... | ..................... | .................... | .................... | .................... |
| AA | 3887 | hCoV-19/Denmark/DCGC-20218/2020\|EPI_ISL_748056\|202 | .................... | .................... | ..................... | .................... | .................... | .................... |
| AA | 3888 | hCoV-19/Denmark/DCGC-20219/2020\|EPI_ISL_748057\|202 | .................... | .................... | ..................... | .................... | .................... | .................... |
| AA | 3889 | hCoV-19/Denmark/DCGC-20220/2020\|EPI_ISL_748058\|202 | .................... | .................... | ..................... | .................... | .................... | .................... |
| AA | 3890 | hCoV-19/Denmark/DCGC-20226/2020\|EPI_ISL_748059\|202 | .................... | .................... | ..................... | .................... | .................... | .................... |
| AA | 3891 | hCoV-19/Denmark/DCGC-20231/2020\|EPI_ISL_748060\|202 | .................... | .................... | ..................... | .................... | .................... | .................... |
| AA | 3892 | hCoV-19/Denmark/DCGC-20232/2020\|EPI_ISL_748061\|202 | .................... | .A.................. | ..................... | .................... | .................... | .................... |
| AA | 3893 | hCoV-19/Denmark/DCGC-20233/2020\|EPI_ISL_748062\|202 | .................... | .................... | ..................... | .................... | .................... | .................... |
| AA | 3894 | hCoV-19/Denmark/DCGC-20234/2020\|EPI_ISL_748063\|202 | .................... | .................... | ..................... | .................... | .................... | .................... |
| AA | 3895 | hCoV-19/Denmark/DCGC-20239/2020\|EPI_ISL_748064\|202 | .................... | .................... | ..................... | .................... | .................... | .................... |
| AA | 3896 | hCoV-19/Denmark/DCGC-20241/2020\|EPI_ISL_748065\|202 | .................... | .................... | ..................... | .................... | .................... | .................... |
| AA | 3897 | hCoV-19/Denmark/DCGC-20242/2020\|EPI_ISL_748066\|202 | .................... | .................... | ..................... | .................... | .................... | .................... |
| AA | 3898 | hCoV-19/Denmark/DCGC-20243/2020\|EPI_ISL_748067\|202 | .................... | .................... | ..................... | .................... | .................... | .................... |
| AA | 3899 | hCoV-19/Denmark/DCGC-20245/2020\|EPI_ISL_748068\|202 | .................... | .................... | ..................... | .................... | .................... | .................... |
| AA | 3900 | hCoV-19/Denmark/DCGC-20246/2020\|EPI_ISL_748069\|202 | .................... | .................... | ..................... | .................... | .................... | .................... |
| AA | 3901 | hCoV-19/Denmark/DCGC-20253/2020\|EPI_ISL_748076\|202 | .................... | .................... | ..................... | .................... | .................... | .................... |
| AA | 3902 | hCoV-19/Denmark/DCGC-20254/2020\|EPI_ISL_748077\|202 | .................... | .................... | ..................... | .................... | .................... | .................... |
| AA | 3903 | hCoV-19/Denmark/DCGC-20258/2020\|EPI_ISL_748080\|202 | .................... | .................... | ..................... | .................... | .................... | .................... |
| AA | 3904 | hCoV-19/Denmark/DCGC-20262/2020\|EPI_ISL_748081\|202 | .................... | .................... | ..................... | .................... | .................... | .................... |
| AA | 3905 | hCoV-19/Denmark/DCGC-20264/2020\|EPI_ISL_748082\|202 | .................... | .................... | ..................... | .................... | .................... | .................... |
| AA | 3906 | hCoV-19/Denmark/DCGC-20265/2020\|EPI_ISL_748083\|202 | .................... | .................... | ..................... | .................... | .................... | .................... |
| AA | 3907 | hCoV-19/Denmark/DCGC-20266/2020\|EPI_ISL_748084\|202 | .................... | .................... | ..................... | .................... | .................... | .................... |
| AA | 3908 | hCoV-19/Denmark/DCGC-20270/2020\|EPI_ISL_748085\|202 | .................... | .................... | ..................... | .................... | .................... | .................... |
| AA | 3909 | hCoV-19/Denmark/DCGC-20273/2020\|EPI_ISL_748086\|202 | .................... | .................... | ..................... | .................... | .................... | .................... |
| AA | 3910 | hCoV-19/Denmark/DCGC-20275/2020\|EPI_ISL_748087\|202 | .................... | .................... | ..................... | .................... | .................... | .................... |
| AA | 3911 | hCoV-19/Denmark/DCGC-20276/2020\|EPI_ISL_748088\|202 | .................... | .................... | ..................... | .................... | .................... | .................... |
| AA | 3912 | hCoV-19/Denmark/DCGC-20278/2020\|EPI_ISL_748089\|202 | .................... | .................... | ..................... | .................... | .................... | .................... |
| AA | 3913 | hCoV-19/Denmark/DCGC-20284/2020\|EPI_ISL_748090\|202 | .................... | .A.................. | ..................... | .................... | .................... | ..A................. |
| AA | 3914 | hCoV-19/Denmark/DCGC-20285/2020\|EPI_ISL_748091\|202 | .................... | .................... | ..................... | .................... | .................... | .................... |
| AA | 3915 | hCoV-19/Denmark/DCGC-20286/2020\|EPI_ISL_748092\|202 | .................... | .................... | ..................... | .................... | .................... | .................... |
| AA | 3916 | hCoV-19/Denmark/DCGC-20287/2020\|EPI_ISL_748093\|202 | .................... | .................... | ..................... | .................... | .................... | .................... |
| AA | 3917 | hCoV-19/Denmark/DCGC-20289/2020\|EPI_ISL_748094\|202 | .................... | .................... | ..................... | .................... | .................... | .................... |
| AA | 3918 | hCoV-19/Denmark/DCGC-20290/2020\|EPI_ISL_748095\|202 | .................... | .................... | ..................... | .................... | .................... | .................... |
| AA | 3919 | hCoV-19/Denmark/DCGC-20291/2020\|EPI_ISL_748096\|202 | .................... | .................... | ..................... | .................... | .................... | .................... |
| AA | 3920 | hCoV-19/Denmark/DCGC-20294/2020\|EPI_ISL_748097\|202 | .................... | .................... | ..................... | .................... | .................... | .................... |
| AA | 3921 | hCoV-19/Denmark/DCGC-20295/2020\|EPI_ISL_748098\|202 | .................... | .................... | ..................... | .................... | .................... | .................... |
| AA | 3922 | hCoV-19/Denmark/DCGC-20296/2020\|EPI_ISL_748099\|202 | .................... | .................... | ..................... | .................... | .................... | .................... |
| AA | 3923 | hCoV-19/Denmark/DCGC-18250/2020\|EPI_ISL_749320\|202 | .................... | .................... | ..................... | .................... | .................... | .................... |

| | | | | | | | | |
|---|---|---|---|---|---|---|---|---|
| AA | 3924 | hCoV-19/Denmark/DCGC-18291/2020\|EPI_ISL_749321\|202 | .................... | .................... | .................... | .................... | .................... | .................... |
| AA | 3925 | hCoV-19/Denmark/DCGC-18292/2020\|EPI_ISL_749322\|202 | .................... | .................... | .................... | .................... | .................... | .................... |
| AA | 3926 | hCoV-19/Denmark/DCGC-18313/2020\|EPI_ISL_749323\|202 | .................... | .................... | .................... | .................... | .................... | .................... |
| AA | 3927 | hCoV-19/Denmark/DCGC-18328/2020\|EPI_ISL_749324\|202 | .................... | .................... | .................... | .................... | .................... | .................... |
| AA | 3928 | hCoV-19/Denmark/DCGC-18374/2020\|EPI_ISL_749325\|202 | .................... | .................... | .................... | .................... | .................... | .................... |
| AA | 3929 | hCoV-19/Denmark/DCGC-18383/2020\|EPI_ISL_749326\|202 | .................... | .................... | .................... | .................... | .................... | .................... |
| AA | 3930 | hCoV-19/Denmark/DCGC-18384/2020\|EPI_ISL_749327\|202 | .................... | .................... | .................... | .................... | .................... | .................... |
| AA | 3931 | hCoV-19/Denmark/DCGC-18392/2020\|EPI_ISL_749328\|202 | .................... | .................... | .................... | .................... | .................... | .................... |
| AA | 3932 | hCoV-19/Denmark/DCGC-18416/2020\|EPI_ISL_749329\|202 | .................... | .................... | .................... | .................... | .................... | .................... |
| AA | 3933 | hCoV-19/Denmark/DCGC-18420/2020\|EPI_ISL_749330\|202 | .................... | .................... | .................... | .................... | .................... | .................... |
| AA | 3934 | hCoV-19/Denmark/DCGC-18456/2020\|EPI_ISL_749331\|202 | .................... | .................... | .................... | .................... | .................... | .................... |
| AA | 3935 | hCoV-19/Denmark/DCGC-18467/2020\|EPI_ISL_749332\|202 | .................... | .................... | .................... | .................... | .................... | .................... |
| AA | 3936 | hCoV-19/Denmark/DCGC-18480/2020\|EPI_ISL_749333\|202 | .................... | .................... | .................... | .................... | .................... | .................... |
| AA | 3937 | hCoV-19/Denmark/DCGC-18494/2020\|EPI_ISL_749334\|202 | .................... | .................... | .................... | .................... | .................... | .................... |
| AA | 3938 | hCoV-19/Denmark/DCGC-18496/2020\|EPI_ISL_749335\|202 | .................... | .................... | .................... | .................... | .................... | .................... |
| AA | 3939 | hCoV-19/Denmark/DCGC-18499/2020\|EPI_ISL_749336\|202 | .................... | .................... | .................... | .................... | .................... | .................... |
| AA | 3940 | hCoV-19/Denmark/DCGC-18551/2020\|EPI_ISL_749337\|202 | .................... | .................... | .................... | .................... | .................... | .................... |
| AA | 3941 | hCoV-19/Denmark/DCGC-18554/2020\|EPI_ISL_749338\|202 | .................... | .................... | .................... | .................... | .................... | .................... |
| AA | 3942 | hCoV-19/Denmark/DCGC-18564/2020\|EPI_ISL_749339\|202 | .................... | .................... | .................... | .................... | .................... | .................... |
| AA | 3943 | hCoV-19/Denmark/DCGC-18585/2020\|EPI_ISL_749340\|202 | .................... | .................... | .................... | .................... | .................... | .................... |
| AA | 3944 | hCoV-19/Denmark/DCGC-18594/2020\|EPI_ISL_749341\|202 | .................... | .................... | .................... | .................... | .................... | .................... |
| AA | 3945 | hCoV-19/Denmark/DCGC-18603/2020\|EPI_ISL_749342\|202 | .................... | .................... | .................... | .................... | .................... | .................... |
| AA | 3946 | hCoV-19/Denmark/DCGC-18605/2020\|EPI_ISL_749343\|202 | .................... | .................... | .................... | .................... | .................... | .................... |
| AA | 3947 | hCoV-19/Denmark/DCGC-18626/2020\|EPI_ISL_749344\|202 | .................... | .................... | .................... | .................... | .................... | .................... |
| AA | 3948 | hCoV-19/Denmark/DCGC-18627/2020\|EPI_ISL_749345\|202 | .................... | .................... | .................... | .................... | .................... | .................... |
| AA | 3949 | hCoV-19/Denmark/DCGC-18635/2020\|EPI_ISL_749346\|202 | .................... | .................... | .................... | .................... | .................... | .................... |
| AA | 3950 | hCoV-19/Denmark/DCGC-18688/2020\|EPI_ISL_749347\|202 | .................... | .................... | .................... | .................... | .................... | .................... |
| AA | 3951 | hCoV-19/Denmark/DCGC-18700/2020\|EPI_ISL_749348\|202 | .................... | .................... | .................... | .................... | .................... | .................... |
| AA | 3952 | hCoV-19/Denmark/DCGC-18706/2020\|EPI_ISL_749349\|202 | .................... | .................... | .................... | .................... | .................... | .................... |
| AA | 3953 | hCoV-19/Denmark/DCGC-17757/2020\|EPI_ISL_749379\|202 | .................... | .................... | .................... | .................... | .................... | .................... |
| AA | 3954 | hCoV-19/Denmark/DCGC-17924/2020\|EPI_ISL_749380\|202 | .................... | .................... | .................... | .................... | .................... | .................... |
| AA | 3955 | hCoV-19/Denmark/DCGC-17957/2020\|EPI_ISL_749381\|202 | .................... | .................... | .................... | .................... | .................... | .................... |
| AA | 3956 | hCoV-19/Denmark/DCGC-18000/2020\|EPI_ISL_749382\|202 | .................... | .................... | .................... | .................... | .................... | .................... |
| AA | 3957 | hCoV-19/Denmark/DCGC-18029/2020\|EPI_ISL_749383\|202 | .................... | .................... | .................... | .................... | .................... | .................... |
| AA | 3958 | hCoV-19/Denmark/DCGC-18040/2020\|EPI_ISL_749384\|202 | .................... | .................... | .................... | .................... | .................... | .................... |
| AA | 3959 | hCoV-19/Denmark/DCGC-18132/2020\|EPI_ISL_749385\|202 | .................... | .................... | .................... | .................... | .................... | .................... |
| AA | 3960 | hCoV-19/Denmark/DCGC-18206/2020\|EPI_ISL_749386\|202 | .................... | .................... | .................... | .................... | .................... | .................... |
| AA | 3961 | hCoV-19/Denmark/DCGC-18207/2020\|EPI_ISL_749387\|202 | .................... | .................... | .................... | .................... | .................... | .................... |
| AA | 3962 | hCoV-19/Denmark/DCGC-18232/2020\|EPI_ISL_749388\|202 | .................... | .................... | .................... | .................... | .................... | .................... |
| AA | 3963 | hCoV-19/Denmark/DCGC-18270/2020\|EPI_ISL_749389\|202 | .................... | .................... | .................... | .................... | .................... | .................... |
| AA | 3964 | hCoV-19/Denmark/DCGC-18324/2020\|EPI_ISL_749390\|202 | .................... | .................... | .................... | .................... | .................... | .................... |
| AA | 3965 | hCoV-19/Denmark/DCGC-18326/2020\|EPI_ISL_749391\|202 | .................... | .................... | .................... | .................... | .................... | .................... |
| AA | 3966 | hCoV-19/Denmark/DCGC-18333/2020\|EPI_ISL_749392\|202 | .................... | .................... | .................... | .................... | .................... | .................... |
| AA | 3967 | hCoV-19/Denmark/DCGC-18339/2020\|EPI_ISL_749393\|202 | .................... | .................... | .................... | .................... | .................... | .................... |
| AA | 3968 | hCoV-19/Denmark/DCGC-18357/2020\|EPI_ISL_749394\|202 | .................... | .................... | .................... | .................... | .................... | .................... |
| AA | 3969 | hCoV-19/Denmark/DCGC-18375/2020\|EPI_ISL_749395\|202 | .................... | .................... | .................... | .................... | .................... | .................... |
| AA | 3970 | hCoV-19/Denmark/DCGC-18457/2020\|EPI_ISL_749396\|202 | .................... | .................... | .................... | .................... | .................... | .................... |
| AA | 3971 | hCoV-19/Denmark/DCGC-18469/2020\|EPI_ISL_749397\|202 | .................... | .................... | .................... | .................... | .................... | .................... |
| AA | 3972 | hCoV-19/Denmark/DCGC-18470/2020\|EPI_ISL_749398\|202 | .................... | .................... | .................... | .................... | .................... | .................... |
| AA | 3973 | hCoV-19/Denmark/DCGC-18487/2020\|EPI_ISL_749399\|202 | .................... | .................... | .................... | .................... | .................... | .................... |
| AA | 3974 | hCoV-19/Denmark/DCGC-16781/2020\|EPI_ISL_750300\|202 | .................... | .................... | .................... | .................... | .................... | .................... |
| AA | 3975 | hCoV-19/Denmark/DCGC-17272/2020\|EPI_ISL_750345\|202 | .................... | .................... | .................... | .................... | .................... | .................... |
| AA | 3976 | hCoV-19/Denmark/DCGC-17387/2020\|EPI_ISL_750346\|202 | .................... | .................... | .................... | .................... | .................... | .................... |
| AA | 3977 | hCoV-19/Denmark/DCGC-17396/2020\|EPI_ISL_750347\|202 | .................... | .................... | .................... | .................... | .................... | .................... |
| AA | 3978 | hCoV-19/Denmark/DCGC-17403/2020\|EPI_ISL_750348\|202 | .................... | .................... | .................... | .................... | .................... | .................... |
| AA | 3979 | hCoV-19/Denmark/DCGC-17404/2020\|EPI_ISL_750349\|202 | .................... | .................... | .................... | .................... | .................... | .................... |
| AA | 3980 | hCoV-19/Denmark/DCGC-17407/2020\|EPI_ISL_750350\|202 | .................... | .................... | .................... | .................... | .................... | .................... |
| AA | 3981 | hCoV-19/Denmark/DCGC-17408/2020\|EPI_ISL_750351\|202 | .................... | .................... | .................... | .................... | .................... | .................... |
| AA | 3982 | hCoV-19/Denmark/DCGC-17424/2020\|EPI_ISL_750352\|202 | .................... | .................... | .................... | .................... | .................... | .................... |
| AA | 3983 | hCoV-19/Denmark/DCGC-17436/2020\|EPI_ISL_750353\|202 | .................... | .................... | .................... | .................... | .................... | .................... |
| AA | 3984 | hCoV-19/Denmark/DCGC-17460/2020\|EPI_ISL_750354\|202 | .................... | .................... | .................... | .................... | .................... | .................... |
| AA | 3985 | hCoV-19/Denmark/DCGC-17466/2020\|EPI_ISL_750355\|202 | .................... | .................... | .................... | .................... | .................... | .................... |
| AA | 3986 | hCoV-19/Denmark/DCGC-17489/2020\|EPI_ISL_750356\|202 | .................... | .................... | .................... | .................... | .................... | .................... |
| AA | 3987 | hCoV-19/Denmark/DCGC-17499/2020\|EPI_ISL_750357\|202 | .................... | .................... | .................... | .................... | .................... | .................... |
| AA | 3988 | hCoV-19/Denmark/DCGC-17512/2020\|EPI_ISL_750358\|202 | .................... | .................... | .................... | .................... | .................... | .................... |
| AA | 3989 | hCoV-19/Denmark/DCGC-17517/2020\|EPI_ISL_750359\|202 | .................... | .................... | .................... | .................... | .................... | .................... |
| AA | 3990 | hCoV-19/Denmark/DCGC-17551/2020\|EPI_ISL_750360\|202 | .................... | .................... | .................... | .................... | .................... | .................... |
| AA | 3991 | hCoV-19/Denmark/DCGC-17610/2020\|EPI_ISL_750361\|202 | .................... | .................... | .................... | .................... | .................... | .................... |
| AA | 3992 | hCoV-19/Denmark/DCGC-17623/2020\|EPI_ISL_750362\|202 | .................... | .................... | .................... | .................... | .................... | .................... |
| AA | 3993 | hCoV-19/Denmark/DCGC-17637/2020\|EPI_ISL_750363\|202 | .................... | .................... | .................... | .................... | .................... | .................... |
| AA | 3994 | hCoV-19/Denmark/DCGC-17640/2020\|EPI_ISL_750364\|202 | .................... | .................... | .................... | .................... | .................... | .................... |
| AA | 3995 | hCoV-19/Denmark/DCGC-17670/2020\|EPI_ISL_750365\|202 | .................... | .................... | .................... | .................... | .................... | .................... |
| AA | 3996 | hCoV-19/Denmark/DCGC-17707/2020\|EPI_ISL_750366\|202 | .................... | .................... | .................... | .................... | .................... | .................... |
| AA | 3997 | hCoV-19/Denmark/DCGC-17741/2020\|EPI_ISL_750367\|202 | .................... | .................... | .................... | .................... | .................... | .................... |
| AA | 3998 | hCoV-19/Denmark/DCGC-17562/2020\|EPI_ISL_750368\|202 | .................... | .................... | .................... | .................... | .................... | .................... |
| AA | 3999 | hCoV-19/Denmark/DCGC-16848/2020\|EPI_ISL_750371\|202 | .................... | .................... | .................... | .................... | .................... | .................... |
| AA | 4000 | hCoV-19/Denmark/DCGC-17266/2020\|EPI_ISL_750372\|202 | .................... | .................... | .................... | .................... | .................... | .................... |
| AA | 4001 | hCoV-19/Denmark/DCGC-16891/2020\|EPI_ISL_750393\|202 | .................... | .................... | .................... | .................... | .................... | .................... |
| AA | 4002 | hCoV-19/Denmark/DCGC-17039/2020\|EPI_ISL_750394\|202 | .................... | .................... | .................... | .................... | .................... | .................... |
| AA | 4003 | hCoV-19/Denmark/DCGC-17051/2020\|EPI_ISL_750395\|202 | .................... | .................... | .................... | .................... | .................... | .................... |

| | | | | | | | | |
|---|---|---|---|---|---|---|---|---|
| AA | 4004 | hCoV-19/Denmark/DCGC-17053/2020\|EPI_ISL_750396\|202 | ···················· | ···················· | ···················· | ···················· | ···················· | ···················· |
| AA | 4005 | hCoV-19/Denmark/DCGC-17075/2020\|EPI_ISL_750397\|202 | ···················· | ···················· | ···················· | ···················· | ···················· | ···················· |
| AA | 4006 | hCoV-19/Denmark/DCGC-17085/2020\|EPI_ISL_750398\|202 | ···················· | ···················· | ···················· | ···················· | ···················· | ···················· |
| AA | 4007 | hCoV-19/Denmark/DCGC-17103/2020\|EPI_ISL_750399\|202 | ···················· | ···················· | ···················· | ···················· | ···················· | ···················· |
| AA | 4008 | hCoV-19/USA/IN-CDC-2-3713960/2020\|EPI_ISL_751600\|2 | ···················· | ···················· | ···················· | ···················· | ···················· | ···················· |
| AA | 4009 | hCoV-19/USA/MI-CDC-2-3713962/2020\|EPI_ISL_751601\|2 | ···················· | ···················· | ···················· | ···················· | ···················· | ···················· |
| AA | 4010 | hCoV-19/USA/DE-CDC-2-3713963/2020\|EPI_ISL_751602\|2 | ···················· | ·A·················· | ···················· | ···················· | ···················· | ···················· |
| AA | 4011 | hCoV-19/USA/HI-CDC-2-3713973/2020\|EPI_ISL_751603\|2 | ···················· | ···················· | ···················· | ···················· | ···················· | ···················· |
| AA | 4012 | hCoV-19/USA/MD-CDC-2-3713976/2020\|EPI_ISL_751604\|2 | ···················· | ···················· | ···················· | ···················· | ···················· | ···················· |
| AA | 4013 | hCoV-19/USA/DE-CDC-2-3713979/2020\|EPI_ISL_751605\|2 | ···················· | ·A·················· | ···················· | ···················· | ···················· | ···················· |
| AA | 4014 | hCoV-19/USA/MD-CDC-2-3713984/2020\|EPI_ISL_751606\|2 | ···················· | ···················· | ···················· | ···················· | ···················· | ···················· |
| AA | 4015 | hCoV-19/USA/MD-CDC-2-3714088/2020\|EPI_ISL_751607\|2 | ···················· | ···················· | ···················· | ···················· | ···················· | ···················· |
| AA | 4016 | hCoV-19/USA/NE-CDC-2-3714268/2020\|EPI_ISL_751608\|2 | ···················· | ···················· | ···················· | ···················· | ···················· | ···················· |
| AA | 4017 | hCoV-19/USA/OH-CDC-2-3714299/2020\|EPI_ISL_751609\|2 | ···················· | ···················· | ···················· | ···················· | ···················· | ···················· |
| AA | 4018 | hCoV-19/USA/OH-CDC-2-3714352/2020\|EPI_ISL_751610\|2 | ···················· | ·············A······ | ···················· | ···················· | ···················· | ···················· |
| AA | 4019 | hCoV-19/USA/SC-CDC-2-3714362/2020\|EPI_ISL_751611\|2 | ···················· | ·A·················· | ···················· | ···················· | ···················· | ···················· |
| AA | 4020 | hCoV-19/USA/ID-CDC-2-3714377/2020\|EPI_ISL_751612\|2 | ···················· | ···················· | ···················· | ···················· | ···················· | ···················· |
| AA | 4021 | hCoV-19/USA/OH-CDC-2-3714378/2020\|EPI_ISL_751613\|2 | ···················· | ···················· | ···················· | ···················· | ···················· | ···················· |
| AA | 4022 | hCoV-19/USA/ID-CDC-2-3714380/2020\|EPI_ISL_751614\|2 | ···················· | ···················· | ···················· | ···················· | ···················· | ···················· |
| AA | 4023 | hCoV-19/USA/OR-CDC-2-3714394/2020\|EPI_ISL_751615\|2 | ···················· | ···················· | ···················· | ···················· | ···················· | ···················· |
| AA | 4024 | hCoV-19/USA/OH-CDC-2-3714454/2020\|EPI_ISL_751616\|2 | ···················· | ···················· | ···················· | ···················· | ···················· | ···················· |
| AA | 4025 | hCoV-19/USA/TX-CDC-2-3714456/2020\|EPI_ISL_751617\|2 | ···················· | ···················· | ···················· | ···················· | ···················· | ···················· |
| AA | 4026 | hCoV-19/USA/ID-CDC-2-3714472/2020\|EPI_ISL_751618\|2 | ···················· | ···················· | ···················· | ···················· | ···················· | ···················· |
| AA | 4027 | hCoV-19/USA/WY-CDC-2-3714478/2020\|EPI_ISL_751619\|2 | ···················· | ···················· | ···················· | ···················· | ···················· | ···················· |
| AA | 4028 | hCoV-19/USA/LA-CDC-2-3714575/2020\|EPI_ISL_751620\|2 | ···················· | ···················· | ···················· | ···················· | ···················· | ···················· |
| AA | 4029 | hCoV-19/USA/DE-CDC-2-3713992/2020\|EPI_ISL_751621\|2 | ···················· | ·A·················· | ···················· | ···················· | ···················· | ···················· |
| AA | 4030 | hCoV-19/USA/MI-CDC-2-3713993/2020\|EPI_ISL_751622\|2 | ···················· | ···················· | ···················· | ···················· | ···················· | ···················· |
| AA | 4031 | hCoV-19/USA/DE-CDC-2-3714051/2020\|EPI_ISL_751623\|2 | ···················· | ···················· | ···················· | ···················· | ···················· | ···················· |
| AA | 4032 | hCoV-19/USA/CT-CDC-2-3714065/2020\|EPI_ISL_751624\|2 | ···················· | ···················· | ···················· | ···················· | ···················· | ···················· |
| AA | 4033 | hCoV-19/USA/MD-CDC-2-3714075/2020\|EPI_ISL_751625\|2 | ···················· | ···················· | ···················· | ···················· | ···················· | ···················· |
| AA | 4034 | hCoV-19/USA/IN-CDC-2-3714077/2020\|EPI_ISL_751626\|2 | ···················· | ···················· | ···················· | ···················· | ···················· | ···················· |
| AA | 4035 | hCoV-19/USA/DE-CDC-2-3714079/2020\|EPI_ISL_751627\|2 | ···················· | ·A·················· | ···················· | ···················· | ···················· | ···················· |
| AA | 4036 | hCoV-19/USA/MD-CDC-2-3714081/2020\|EPI_ISL_751628\|2 | ···················· | ·A·················· | ···················· | ···················· | ···················· | ···················· |
| AA | 4037 | hCoV-19/USA/MD-CDC-2-3714084/2020\|EPI_ISL_751629\|2 | ···················· | ···················· | ···················· | ···················· | ···················· | ···················· |
| AA | 4038 | hCoV-19/USA/CO-CDC-2-3714087/2020\|EPI_ISL_751630\|2 | ···················· | ···················· | ···················· | ···················· | ···················· | ···················· |
| AA | 4039 | hCoV-19/USA/DE-CDC-2-3714090/2020\|EPI_ISL_751631\|2 | ···················· | ·A·················· | ···················· | ···················· | ···················· | ···················· |
| AA | 4040 | hCoV-19/USA/IN-CDC-2-3713999/2020\|EPI_ISL_751676\|2 | ···················· | ···················· | ···················· | ···················· | ···················· | ···················· |
| AA | 4041 | hCoV-19/USA/KS-CDC-2-3714060/2020\|EPI_ISL_751677\|2 | ···················· | ···················· | ···················· | ···················· | ···················· | ···················· |
| AA | 4042 | hCoV-19/USA/NH-CDC-2-3714186/2020\|EPI_ISL_751678\|2 | ···················· | ···················· | ···················· | ···················· | ···················· | ···················· |
| AA | 4043 | hCoV-19/USA/MS-CDC-2-3714198/2020\|EPI_ISL_751679\|2 | ···················· | ···················· | ···················· | ···················· | ···················· | ···················· |
| AA | 4044 | hCoV-19/USA/NJ-CDC-2-3714257/2020\|EPI_ISL_751680\|2 | ···················· | ·A·················· | ···················· | ···················· | ···················· | ···················· |
| AA | 4045 | hCoV-19/USA/NE-CDC-2-3714274/2020\|EPI_ISL_751681\|2 | ···················· | ···················· | ···················· | ···················· | ···················· | ···················· |
| AA | 4046 | hCoV-19/USA/NY-CDC-2-3714276/2020\|EPI_ISL_751682\|2 | ···················· | ···················· | ···················· | ···················· | ···················· | ···················· |
| AA | 4047 | hCoV-19/USA/MS-CDC-2-3714288/2020\|EPI_ISL_751683\|2 | ···················· | ·A·················· | ···················· | ···················· | ···················· | ···················· |
| AA | 4048 | hCoV-19/USA/TX-CDC-2-3714291/2020\|EPI_ISL_751684\|2 | ···················· | ···················· | ···················· | ···················· | ···················· | ···················· |
| AA | 4049 | hCoV-19/USA/VA-CDC-2-3714292/2020\|EPI_ISL_751685\|2 | ···················· | ·············A······ | ···················· | ···················· | ···················· | ···················· |
| AA | 4050 | hCoV-19/USA/VA-CDC-2-3714368/2020\|EPI_ISL_751686\|2 | ···················· | ···················· | ···················· | ···················· | ···················· | ···················· |
| AA | 4051 | hCoV-19/USA/IA-CDC-2-3714371/2020\|EPI_ISL_751687\|2 | ···················· | ···················· | ···················· | ···················· | ···················· | ···················· |
| AA | 4052 | hCoV-19/USA/WY-CDC-2-3714460/2020\|EPI_ISL_751688\|2 | ···················· | ···················· | ···················· | ···················· | ···················· | ···················· |
| AA | 4053 | hCoV-19/USA/OR-CDC-2-3714464/2020\|EPI_ISL_751689\|2 | ···················· | ·A·················· | ···················· | ···················· | ···················· | ···················· |
| AA | 4054 | hCoV-19/USA/VA-CDC-2-3714475/2020\|EPI_ISL_751690\|2 | ···················· | ···················· | ···················· | ···················· | ···················· | ···················· |
| AA | 4055 | hCoV-19/USA/WY-CDC-2-3714476/2020\|EPI_ISL_751691\|2 | ···················· | ·A·················· | ···················· | ···················· | ···················· | ···················· |
| AA | 4056 | hCoV-19/USA/IA-CDC-2-3714477/2020\|EPI_ISL_751692\|2 | ···················· | ···················· | ···················· | ···················· | ···················· | ···················· |
| AA | 4057 | hCoV-19/USA/OR-CDC-2-3714482/2020\|EPI_ISL_751693\|2 | ···················· | ·············M······ | ···················· | ···················· | ···················· | ···················· |
| AA | 4058 | hCoV-19/USA/AK-CDC-2-3714492/2020\|EPI_ISL_751694\|2 | ···················· | ···················· | ···················· | ···················· | ···················· | ···················· |
| AA | 4059 | hCoV-19/USA/RI-CDC-2-3714565/2020\|EPI_ISL_751695\|2 | ···················· | ···················· | ···················· | ···················· | ···················· | ···················· |
| AA | 4060 | hCoV-19/USA/LA-CDC-2-3714577/2020\|EPI_ISL_751696\|2 | ···················· | ···················· | ···················· | ···················· | ···················· | ···················· |
| AA | 4061 | hCoV-19/USA/AR-CDC-2-3713953/2020\|EPI_ISL_751697\|2 | ···················· | ···················· | ···················· | ···················· | ···················· | ···················· |
| AA | 4062 | hCoV-19/USA/AR-CDC-2-3713965/2020\|EPI_ISL_751698\|2 | ···················· | ···················· | ···················· | ···················· | ···················· | ···················· |
| AA | 4063 | hCoV-19/USA/AR-CDC-2-3713967/2020\|EPI_ISL_751699\|2 | ···················· | ···················· | ···················· | ···················· | ···················· | ···················· |
| AA | 4064 | hCoV-19/USA/HI-H200383/2020\|EPI_ISL_752930\|2020-09 | ···················· | ·A·················· | ···················· | ···················· | ···················· | ···················· |
| AA | 4065 | hCoV-19/USA/HI-H200384/2020\|EPI_ISL_752931\|2020-09 | ···················· | ·A·················· | ···················· | ···················· | ···················· | ···················· |
| AA | 4066 | hCoV-19/USA/HI-H200385/2020\|EPI_ISL_752932\|2020-09 | ···················· | ·A·················· | ···················· | ···················· | ···················· | ···················· |
| AA | 4067 | hCoV-19/USA/HI-H200386/2020\|EPI_ISL_752933\|2020-09 | ···················· | ·A·················· | ···················· | ···················· | ···················· | ···················· |
| AA | 4068 | hCoV-19/USA/HI-H200387/2020\|EPI_ISL_752934\|2020-09 | ···················· | ·A·················· | ···················· | ···················· | ···················· | ···················· |
| AA | 4069 | hCoV-19/USA/HI-H200388/2020\|EPI_ISL_752935\|2020-09 | ···················· | ·A·················· | ···················· | ···················· | ···················· | ···················· |
| AA | 4070 | hCoV-19/USA/HI-H200389/2020\|EPI_ISL_752936\|2020-09 | ···················· | ·A·················· | ···················· | ···················· | ···················· | ···················· |
| AA | 4071 | hCoV-19/USA/HI-H200390/2020\|EPI_ISL_752937\|2020-09 | ···················· | ·A·················· | ···················· | ···················· | ···················· | ···················· |
| AA | 4072 | hCoV-19/USA/HI-H200391/2020\|EPI_ISL_752938\|2020-09 | ···················· | ·A·················· | ···················· | ···················· | ···················· | ···················· |
| AA | 4073 | hCoV-19/USA/HI-H200392/2020\|EPI_ISL_752939\|2020-09 | ···················· | ·A·················· | ···················· | ···················· | ···················· | ···················· |
| AA | 4074 | hCoV-19/USA/HI-H200393/2020\|EPI_ISL_752940\|2020-09 | ···················· | ·A·················· | ···················· | ···················· | ···················· | ···················· |
| AA | 4075 | hCoV-19/USA/HI-H200395/2020\|EPI_ISL_752941\|2020-09 | ···················· | ·A·················· | ···················· | ···················· | ···················· | ···················· |
| AA | 4076 | hCoV-19/USA/HI-H200396/2020\|EPI_ISL_752942\|2020-09 | ···················· | ·A·················· | ···················· | ···················· | ···················· | ···················· |
| AA | 4077 | hCoV-19/USA/HI-H200398/2020\|EPI_ISL_752943\|2020-09 | ···················· | ·A·················· | ···················· | ···················· | ···················· | ···················· |
| AA | 4078 | hCoV-19/USA/HI-H200399/2020\|EPI_ISL_752944\|2020-09 | ···················· | ·A·················· | ···················· | ···················· | ···················· | ···················· |
| AA | 4079 | hCoV-19/USA/HI-H200400/2020\|EPI_ISL_752945\|2020-09 | ···················· | ·A·················· | ···················· | ···················· | ···················· | ···················· |
| AA | 4080 | hCoV-19/USA/HI-H200401/2020\|EPI_ISL_752946\|2020-09 | ···················· | ·A·················· | ···················· | ···················· | ···················· | ···················· |
| AA | 4081 | hCoV-19/USA/HI-H200402/2020\|EPI_ISL_752947\|2020-09 | ···················· | ·A·················· | ···················· | ···················· | ···················· | ···················· |
| AA | 4082 | hCoV-19/USA/HI-H200403/2020\|EPI_ISL_752948\|2020-09 | ···················· | ·A·················· | ···················· | ···················· | ···················· | ···················· |
| AA | 4083 | hCoV-19/USA/HI-H200404/2020\|EPI_ISL_752949\|2020-09 | ···················· | ·A·················· | ···················· | ···················· | ···················· | ···················· |

| | | | | | | | |
|---|---|---|---|---|---|---|---|
| AA | 4084 hCoV-19/USA/HI-H200405/2020\|EPI_ISL_752950\|2020-07 | ···················· | ···················· | ····················· | ···················· | ···················· | ···················· |
| AA | 4085 hCoV-19/USA/HI-H200406/2020\|EPI_ISL_752951\|2020-07 | ···················· | ·A·················· | ····················· | ···················· | ···················· | ···················· |
| AA | 4086 hCoV-19/USA/HI-H200407/2020\|EPI_ISL_752952\|2020-07 | ···················· | ·A·················· | ····················· | ···················· | ···················· | ···················· |
| AA | 4087 hCoV-19/USA/HI-H200408/2020\|EPI_ISL_752953\|2020-08 | ···················· | ·A·················· | ····················· | ···················· | ···················· | ···················· |
| AA | 4088 hCoV-19/USA/HI-H200409/2020\|EPI_ISL_752954\|2020-08 | ···················· | ·A·················· | ····················· | ···················· | ···················· | ···················· |
| AA | 4089 hCoV-19/USA/HI-H200411/2020\|EPI_ISL_752955\|2020-08 | ···················· | ·A·················· | ····················· | ···················· | ···················· | ···················· |
| AA | 4090 hCoV-19/USA/HI-H200412/2020\|EPI_ISL_752956\|2020-08 | ···················· | ·A·················· | ····················· | ···················· | ···················· | ···················· |
| AA | 4091 hCoV-19/USA/HI-H200413/2020\|EPI_ISL_752957\|2020-08 | ···················· | ···················· | ····················· | ···················· | ···················· | ···················· |
| AA | 4092 hCoV-19/USA/HI-H200414/2020\|EPI_ISL_752958\|2020-08 | ···················· | ·A·················· | ····················· | ···················· | ···················· | ···················· |
| AA | 4093 hCoV-19/USA/HI-H200415/2020\|EPI_ISL_752959\|2020-08 | ···················· | ·A·················· | ····················· | ···················· | ···················· | ···················· |
| AA | 4094 hCoV-19/USA/HI-H200416/2020\|EPI_ISL_752960\|2020-08 | ···················· | ···················· | ····················· | ···················· | ···················· | ···················· |
| AA | 4095 hCoV-19/USA/HI-H200417/2020\|EPI_ISL_752961\|2020-08 | ···················· | ·A·················· | ····················· | ···················· | ···················· | ···················· |
| AA | 4096 hCoV-19/USA/HI-H200418/2020\|EPI_ISL_752962\|2020-08 | ···················· | ·A·················· | ····················· | ···················· | ···················· | ···················· |
| AA | 4097 hCoV-19/Denmark/DCGC-20247/2020\|EPI_ISL_748070\|202 | ···················· | ···················· | ····················· | ···················· | ···················· | ···················· |
| AA | 4098 hCoV-19/Denmark/DCGC-20248/2020\|EPI_ISL_748071\|202 | ···················· | ···················· | ····················· | ···················· | ···················· | ···················· |
| AA | 4099 hCoV-19/Denmark/DCGC-20249/2020\|EPI_ISL_748072\|202 | ···················· | ···················· | ····················· | ···················· | ···················· | ···················· |
| AA | 4100 hCoV-19/Denmark/DCGC-20250/2020\|EPI_ISL_748073\|202 | ···················· | ···················· | ····················· | ···················· | ···················· | ···················· |
| AA | 4101 hCoV-19/Denmark/DCGC-20251/2020\|EPI_ISL_748074\|202 | ···················· | ·A·················· | ····················· | ···················· | ···················· | ···················· |
| AA | 4102 hCoV-19/Denmark/DCGC-20252/2020\|EPI_ISL_748075\|202 | ···················· | ···················· | ····················· | ···················· | ···················· | ···················· |
| AA | 4103 hCoV-19/Denmark/DCGC-20255/2020\|EPI_ISL_748078\|202 | ···················· | ···················· | ····················· | ···················· | ······T············· | ···················· |
| AA | 4104 hCoV-19/Denmark/DCGC-20257/2020\|EPI_ISL_748079\|202 | ···················· | ·A·················· | ····················· | ···················· | ···················· | ···················· |
| AA | 4105 hCoV-19/Denmark/DCGC-20297/2020\|EPI_ISL_748100\|202 | ···················· | ···················· | ····················· | ···················· | ···················· | ···················· |
| AA | 4106 hCoV-19/Denmark/DCGC-20298/2020\|EPI_ISL_748101\|202 | ···················· | ···················· | ····················· | ···················· | ···················· | ···················· |
| AA | 4107 hCoV-19/Denmark/DCGC-20300/2020\|EPI_ISL_748102\|202 | ···················· | ···················· | ····················· | ···················· | ···················· | ···················· |
| AA | 4108 hCoV-19/Denmark/DCGC-20302/2020\|EPI_ISL_748103\|202 | ···················· | ···················· | ····················· | ···················· | ···················· | ···················· |
| AA | 4109 hCoV-19/Denmark/DCGC-20306/2020\|EPI_ISL_748104\|202 | ···················· | ···················· | ····················· | ···················· | ···················· | ···················· |
| AA | 4110 hCoV-19/Denmark/DCGC-20307/2020\|EPI_ISL_748105\|202 | ···················· | ···················· | ····················· | ···················· | ···················· | ···················· |
| AA | 4111 hCoV-19/Denmark/DCGC-20308/2020\|EPI_ISL_748106\|202 | ···················· | ···················· | ····················· | ···················· | ···················· | ···················· |
| AA | 4112 hCoV-19/Denmark/DCGC-20310/2020\|EPI_ISL_748107\|202 | ···················· | ···················· | ····················· | ···················· | ···················· | ···················· |
| AA | 4113 hCoV-19/Denmark/DCGC-20314/2020\|EPI_ISL_748108\|202 | ···················· | ···················· | ····················· | ···················· | ···················· | ···················· |
| AA | 4114 hCoV-19/Denmark/DCGC-20316/2020\|EPI_ISL_748109\|202 | ···················· | ·A·················· | ····················· | ···················· | ···················· | ···A················ |
| AA | 4115 hCoV-19/Denmark/DCGC-17802/2020\|EPI_ISL_749410\|202 | ···················· | ···················· | ····················· | ···················· | ···················· | ···················· |
| AA | 4116 hCoV-19/Denmark/DCGC-17819/2020\|EPI_ISL_749411\|202 | ···················· | ···················· | ····················· | ···················· | ···················· | ···················· |
| AA | 4117 hCoV-19/Denmark/DCGC-17825/2020\|EPI_ISL_749412\|202 | ···················· | ···················· | ····················· | ···················· | ···················· | ···················· |
| AA | 4118 hCoV-19/Denmark/DCGC-17833/2020\|EPI_ISL_749413\|202 | ···················· | ···················· | ····················· | ···················· | ···················· | ···················· |
| AA | 4119 hCoV-19/Denmark/DCGC-17844/2020\|EPI_ISL_749414\|202 | ···················· | ···················· | ····················· | ···················· | ···················· | ···················· |
| AA | 4120 hCoV-19/Denmark/DCGC-17853/2020\|EPI_ISL_749415\|202 | ···················· | ···················· | ····················· | ···················· | ···················· | ···················· |
| AA | 4121 hCoV-19/Denmark/DCGC-17854/2020\|EPI_ISL_749416\|202 | ···················· | ···················· | ····················· | ···················· | ···················· | ···················· |
| AA | 4122 hCoV-19/Denmark/DCGC-17870/2020\|EPI_ISL_749417\|202 | ···················· | ···················· | ····················· | ···················· | ···················· | ···················· |
| AA | 4123 hCoV-19/Denmark/DCGC-17887/2020\|EPI_ISL_749418\|202 | ···················· | ···················· | ····················· | ···················· | ···················· | ···················· |
| AA | 4124 hCoV-19/Denmark/DCGC-17890/2020\|EPI_ISL_749419\|202 | ···················· | ···················· | ····················· | ···················· | ···················· | ···················· |
| AA | 4125 hCoV-19/Denmark/DCGC-17892/2020\|EPI_ISL_749420\|202 | ···················· | ···················· | ····················· | ···················· | ···················· | ···················· |
| AA | 4126 hCoV-19/Denmark/DCGC-17910/2020\|EPI_ISL_749421\|202 | ···················· | ···················· | ····················· | ···················· | ···················· | ···················· |
| AA | 4127 hCoV-19/Denmark/DCGC-17941/2020\|EPI_ISL_749422\|202 | ···················· | ···················· | ····················· | ···················· | ···················· | ···················· |
| AA | 4128 hCoV-19/Denmark/DCGC-17946/2020\|EPI_ISL_749423\|202 | ···················· | ···················· | ····················· | ···················· | ···················· | ···················· |
| AA | 4129 hCoV-19/Denmark/DCGC-17956/2020\|EPI_ISL_749424\|202 | ···················· | ···················· | ····················· | ···················· | ···················· | ···················· |
| AA | 4130 hCoV-19/Denmark/DCGC-17960/2020\|EPI_ISL_749425\|202 | ···················· | ···················· | ····················· | ···················· | ···················· | ···················· |
| AA | 4131 hCoV-19/Denmark/DCGC-17967/2020\|EPI_ISL_749426\|202 | ···················· | ···················· | ····················· | ···················· | ···················· | ···················· |
| AA | 4132 hCoV-19/Denmark/DCGC-17968/2020\|EPI_ISL_749427\|202 | ···················· | ···················· | ····················· | ···················· | ···················· | ···················· |
| AA | 4133 hCoV-19/Denmark/DCGC-17973/2020\|EPI_ISL_749428\|202 | ···················· | ···················· | ····················· | ···················· | ···················· | ···················· |
| AA | 4134 hCoV-19/Denmark/DCGC-18013/2020\|EPI_ISL_749429\|202 | ···················· | ···················· | ····················· | ···················· | ···················· | ···················· |
| AA | 4135 hCoV-19/Denmark/DCGC-18014/2020\|EPI_ISL_749430\|202 | ···················· | ···················· | ····················· | ···················· | ···················· | ···················· |
| AA | 4136 hCoV-19/Denmark/DCGC-18024/2020\|EPI_ISL_749431\|202 | ···················· | ···················· | ····················· | ···················· | ···················· | ···················· |
| AA | 4137 hCoV-19/Denmark/DCGC-18025/2020\|EPI_ISL_749432\|202 | ···················· | ···················· | ····················· | ···················· | ···················· | ···················· |
| AA | 4138 hCoV-19/Denmark/DCGC-18070/2020\|EPI_ISL_749433\|202 | ···················· | ···················· | ····················· | ···················· | ···················· | ···················· |
| AA | 4139 hCoV-19/Denmark/DCGC-18073/2020\|EPI_ISL_749434\|202 | ···················· | ···················· | ····················· | ···················· | ···················· | ···················· |
| AA | 4140 hCoV-19/Denmark/DCGC-18092/2020\|EPI_ISL_749435\|202 | ···················· | ···················· | ····················· | ···················· | ···C················ | ···················· |
| AA | 4141 hCoV-19/Denmark/DCGC-18094/2020\|EPI_ISL_749436\|202 | ···················· | ···················· | ····················· | ···················· | ···················· | ···················· |
| AA | 4142 hCoV-19/Denmark/DCGC-18101/2020\|EPI_ISL_749437\|202 | ···················· | ···················· | ····················· | ···················· | ···················· | ···················· |
| AA | 4143 hCoV-19/Denmark/DCGC-18104/2020\|EPI_ISL_749438\|202 | ···················· | ···················· | ····················· | ···················· | ···················· | ···················· |
| AA | 4144 hCoV-19/Denmark/DCGC-18121/2020\|EPI_ISL_749439\|202 | ···················· | ···················· | ····················· | ···················· | ···················· | ···················· |
| AA | 4145 hCoV-19/Denmark/DCGC-16950/2020\|EPI_ISL_750422\|202 | ···················· | ···················· | ····················· | ···················· | ····T··············· | ···················· |
| AA | 4146 hCoV-19/Denmark/DCGC-16955/2020\|EPI_ISL_750423\|202 | ···················· | ···················· | ····················· | ···················· | ···················· | ···················· |
| AA | 4147 hCoV-19/Denmark/DCGC-16958/2020\|EPI_ISL_750424\|202 | ···················· | ···················· | ····················· | ···················· | ···················· | ···················· |
| AA | 4148 hCoV-19/Denmark/DCGC-16976/2020\|EPI_ISL_750425\|202 | ···················· | ···················· | ····················· | ···················· | ···················· | ···················· |
| AA | 4149 hCoV-19/Denmark/DCGC-16995/2020\|EPI_ISL_750426\|202 | ···················· | ···················· | ····················· | ···················· | ···················· | ···················· |
| AA | 4150 hCoV-19/Denmark/DCGC-17003/2020\|EPI_ISL_750427\|202 | ···················· | ···················· | ····················· | ···················· | ···················· | ···················· |
| AA | 4151 hCoV-19/Denmark/DCGC-17025/2020\|EPI_ISL_750428\|202 | ···················· | ···················· | ····················· | ···················· | ···················· | ···················· |
| AA | 4152 hCoV-19/Denmark/DCGC-17030/2020\|EPI_ISL_750429\|202 | ···················· | ···················· | ····················· | ···················· | ···················· | ···················· |
| AA | 4153 hCoV-19/Uruguay/TYT-M133/2020\|EPI_ISL_750430\|2020- | ···················· | ···················· | ····················· | ···················· | ···················· | ···················· |
| AA | 4154 hCoV-19/Denmark/DCGC-17044/2020\|EPI_ISL_750431\|202 | ···················· | ···················· | ····················· | ···················· | ···················· | ···················· |
| AA | 4155 hCoV-19/Denmark/DCGC-17055/2020\|EPI_ISL_750432\|202 | ···················· | ···················· | ····················· | ···················· | ···················· | ···················· |
| AA | 4156 hCoV-19/Denmark/DCGC-17058/2020\|EPI_ISL_750433\|202 | ···················· | ···················· | ····················· | ···················· | ···················· | ···················· |
| AA | 4157 hCoV-19/Denmark/DCGC-17074/2020\|EPI_ISL_750434\|202 | ···················· | ···················· | ····················· | ···················· | ···················· | ···················· |
| AA | 4158 hCoV-19/Denmark/DCGC-17078/2020\|EPI_ISL_750435\|202 | ···················· | ···················· | ····················· | ···················· | ···················· | ···················· |
| AA | 4159 hCoV-19/Denmark/DCGC-17093/2020\|EPI_ISL_750436\|202 | ···················· | ···················· | ····················· | ···················· | ···················· | ···················· |
| AA | 4160 hCoV-19/Denmark/DCGC-17096/2020\|EPI_ISL_750437\|202 | ···················· | ···················· | ····················· | ···················· | ···················· | ···················· |
| AA | 4161 hCoV-19/Denmark/DCGC-17102/2020\|EPI_ISL_750438\|202 | ···················· | ···················· | ····················· | ···················· | ···················· | ···················· |
| AA | 4162 hCoV-19/Denmark/DCGC-17110/2020\|EPI_ISL_750439\|202 | ···················· | ···················· | ····················· | ···················· | ···················· | ···················· |
| AA | 4163 hCoV-19/Denmark/DCGC-17149/2020\|EPI_ISL_750440\|202 | ···················· | ···················· | ····················· | ···················· | ···················· | ···················· |

| | | | | | | | | | |
|---|---|---|---|---|---|---|---|---|---|
| AA | 4164 | hCoV-19/Denmark/DCGC-17161/2020\|EPI_ISL_750441\|202 | ···················· | ···················· | ···················· | ···················· | ···················· | ···················· | |
| AA | 4165 | hCoV-19/Denmark/DCGC-17167/2020\|EPI_ISL_750442\|202 | ···················· | ···················· | ···················· | ···················· | ···················· | ···················· | |
| AA | 4166 | hCoV-19/Denmark/DCGC-17186/2020\|EPI_ISL_750443\|202 | ···················· | ···················· | ···················· | ···················· | ····T··············· | ···················· | |
| AA | 4167 | hCoV-19/Denmark/DCGC-17214/2020\|EPI_ISL_750444\|202 | ···················· | ···················· | ···················· | ···················· | ···················· | ···················· | |
| AA | 4168 | hCoV-19/Denmark/DCGC-17247/2020\|EPI_ISL_750445\|202 | ···················· | ···················· | ···················· | ···················· | ···················· | ···················· | |
| AA | 4169 | hCoV-19/Denmark/DCGC-17256/2020\|EPI_ISL_750446\|202 | ···················· | ···················· | ···················· | ···················· | ···················· | ···················· | |
| AA | 4170 | hCoV-19/Denmark/DCGC-17258/2020\|EPI_ISL_750447\|202 | ···················· | ···················· | ···················· | ···················· | ···················· | ···················· | |
| AA | 4171 | hCoV-19/Denmark/DCGC-17286/2020\|EPI_ISL_750448\|202 | ···················· | ···················· | ···················· | ···················· | ···················· | ···················· | |
| AA | 4172 | hCoV-19/Denmark/DCGC-17288/2020\|EPI_ISL_750449\|202 | ···················· | ···················· | ···················· | ···················· | ···················· | ···················· | |
| AA | 4173 | hCoV-19/Denmark/DCGC-17295/2020\|EPI_ISL_750451\|202 | ···················· | ···················· | ···················· | ···················· | ···················· | ···················· | |
| AA | 4174 | hCoV-19/Denmark/DCGC-17339/2020\|EPI_ISL_750453\|202 | ···················· | ···················· | ···················· | ···················· | ···················· | ···················· | |
| AA | 4175 | hCoV-19/Denmark/DCGC-17345/2020\|EPI_ISL_750454\|202 | ···················· | ···················· | ···················· | ···················· | ···················· | ···················· | |
| AA | 4176 | hCoV-19/USA/MD-CDC-2-3713990/2020\|EPI_ISL_751700\|2 | ···················· | ···················· | ···················· | ···················· | ···················· | ···················· | |
| AA | 4177 | hCoV-19/USA/CO-CDC-2-3713995/2020\|EPI_ISL_751701\|2 | ···················· | ···················· | ···················· | ···················· | ···················· | ···················· | |
| AA | 4178 | hCoV-19/USA/MD-CDC-2-3713997/2020\|EPI_ISL_751702\|2 | ···················· | ···················· | ···················· | ···················· | ···················· | ···················· | |
| AA | 4179 | hCoV-19/USA/MD-CDC-2-3713998/2020\|EPI_ISL_751703\|2 | ···················· | ·A·················· | ···················· | ···················· | ···················· | ···················· | |
| AA | 4180 | hCoV-19/USA/CO-CDC-2-3714058/2020\|EPI_ISL_751704\|2 | ···················· | ···················· | ···················· | ···················· | ···················· | ···················· | |
| AA | 4181 | hCoV-19/USA/CO-CDC-2-3714063/2020\|EPI_ISL_751705\|2 | ···················· | ·A·················· | ···················· | ···················· | ···················· | ···················· | |
| AA | 4182 | hCoV-19/USA/CT-CDC-2-3714083/2020\|EPI_ISL_751706\|2 | ···················· | ···················· | ···················· | ···················· | ···················· | ···················· | |
| AA | 4183 | hCoV-19/USA/KS-CDC-2-3714085/2020\|EPI_ISL_751707\|2 | ···················· | ···················· | ···················· | ···················· | ···················· | ···················· | |
| AA | 4184 | hCoV-19/USA/NV-CDC-2-3714100/2020\|EPI_ISL_751708\|2 | ···················· | ·A·················· | ···················· | ···················· | ···················· | ···················· | |
| AA | 4185 | hCoV-19/USA/NJ-CDC-2-3714156/2020\|EPI_ISL_751709\|2 | ···················· | ·A·················· | ···················· | ···················· | ···················· | ···················· | |
| AA | 4186 | hCoV-19/USA/AL-CDC-2-3714566/2020\|EPI_ISL_751753\|2 | ···················· | ···················· | ···················· | ···················· | ···················· | ···················· | |
| AA | 4187 | hCoV-19/USA/KY-CDC-2-3714582/2020\|EPI_ISL_751754\|2 | ···················· | ···················· | ···················· | ···················· | ···················· | ···················· | |
| AA | 4188 | hCoV-19/USA/AL-CDC-2-3714585/2020\|EPI_ISL_751755\|2 | ···················· | ·A·················· | ···················· | ···················· | ···················· | ···················· | |
| AA | 4189 | hCoV-19/USA/AL-CDC-2-3714586/2020\|EPI_ISL_751756\|2 | ···················· | ·A·················· | ···················· | ···················· | ···················· | ···················· | |
| AA | 4190 | hCoV-19/USA/RI-CDC-2-3714591/2020\|EPI_ISL_751757\|2 | ···················· | ···················· | ···················· | ···················· | ···················· | ···················· | |
| AA | 4191 | hCoV-19/USA/HI-CDC-2-3713954/2020\|EPI_ISL_751758\|2 | ···················· | ···················· | ···················· | ···················· | ···················· | ···················· | |
| AA | 4192 | hCoV-19/USA/CT-CDC-2-3713964/2020\|EPI_ISL_751759\|2 | ···················· | ···················· | ···················· | ···················· | ···················· | ···················· | |
| AA | 4193 | hCoV-19/USA/AR-CDC-2-3713966/2020\|EPI_ISL_751760\|2 | ···················· | ···················· | ···················· | ···················· | ···················· | ···················· | |
| AA | 4194 | hCoV-19/USA/HI-CDC-2-3713972/2020\|EPI_ISL_751761\|2 | ···················· | ·A·················· | ···················· | ···················· | ···················· | ···················· | |
| AA | 4195 | hCoV-19/USA/CT-CDC-2-3713974/2020\|EPI_ISL_751762\|2 | ···················· | ···················· | ···················· | ···················· | ···················· | ···················· | |
| AA | 4196 | hCoV-19/USA/CO-CDC-2-3713975/2020\|EPI_ISL_751763\|2 | ···················· | ···················· | ···················· | ···················· | ···················· | ···················· | |
| AA | 4197 | hCoV-19/USA/AR-CDC-2-3713989/2020\|EPI_ISL_751764\|2 | ···················· | ···················· | ···················· | ···················· | ···················· | ···················· | |
| AA | 4198 | hCoV-19/USA/AR-CDC-2-3714000/2020\|EPI_ISL_751765\|2 | ···················· | ···················· | ···················· | ···················· | ···················· | ···················· | |
| AA | 4199 | hCoV-19/USA/CO-CDC-2-3714064/2020\|EPI_ISL_751766\|2 | ···················· | ···················· | ···················· | ···················· | ···················· | ···················· | |
| AA | 4200 | hCoV-19/USA/HI-CDC-2-3714066/2020\|EPI_ISL_751767\|2 | ···················· | ···················· | ···················· | ···················· | ···················· | ···················· | |
| AA | 4201 | hCoV-19/USA/IN-CDC-2-3714067/2020\|EPI_ISL_751768\|2 | ···················· | ···················· | ···················· | ···················· | ···················· | ···················· | |
| AA | 4202 | hCoV-19/USA/MI-CDC-2-3714072/2020\|EPI_ISL_751769\|2 | ···················· | ···················· | ···················· | ···················· | ···················· | ···················· | |
| AA | 4203 | hCoV-19/USA/IN-CDC-2-3714078/2020\|EPI_ISL_751770\|2 | ···················· | ·A·················· | ···················· | ···················· | ···················· | ···················· | |
| AA | 4204 | hCoV-19/USA/MD-CDC-2-3714082/2020\|EPI_ISL_751771\|2 | ···················· | ·A·················· | ···················· | ···················· | ···················· | ···················· | |
| AA | 4205 | hCoV-19/USA/HI-CDC-2-3714093/2020\|EPI_ISL_751772\|2 | ···················· | ···················· | ···················· | ···················· | ···················· | ···················· | |
| AA | 4206 | hCoV-19/USA/NJ-CDC-2-3714161/2020\|EPI_ISL_751773\|2 | ···················· | ·A·················· | ···················· | ···················· | ···················· | ···················· | |
| AA | 4207 | hCoV-19/USA/MO-CDC-2-3714163/2020\|EPI_ISL_751774\|2 | ···················· | ···················· | ···················· | ···················· | ···················· | ···················· | |
| AA | 4208 | hCoV-19/USA/MT-CDC-2-3714190/2020\|EPI_ISL_751775\|2 | ···················· | ···················· | ···················· | ···················· | ···················· | ···················· | |
| AA | 4209 | hCoV-19/USA/MS-CDC-2-3714197/2020\|EPI_ISL_751776\|2 | ···················· | ···················· | ···················· | ···················· | ···················· | ···················· | |
| AA | 4210 | hCoV-19/USA/OH-CDC-2-3714258/2020\|EPI_ISL_751777\|2 | ···················· | ·A·················· | ···················· | ···················· | ···················· | ···················· | |
| AA | 4211 | hCoV-19/USA/NV-CDC-2-3714281/2020\|EPI_ISL_751778\|2 | ···················· | ···················· | ···················· | ···················· | ···················· | ···················· | |
| AA | 4212 | hCoV-19/USA/IA-CDC-2-3714295/2020\|EPI_ISL_751779\|2 | ···················· | ···················· | ···················· | ···················· | ···················· | ···················· | |
| AA | 4213 | hCoV-19/USA/ID-CDC-2-3714366/2020\|EPI_ISL_751782\|2 | ···················· | ···················· | ···················· | ···················· | ···················· | ···················· | |
| AA | 4214 | hCoV-19/USA/IA-CDC-2-3714367/2020\|EPI_ISL_751783\|2 | ···················· | ···················· | ···················· | ···················· | ···················· | ···················· | |
| AA | 4215 | hCoV-19/USA/VA-CDC-2-3714369/2020\|EPI_ISL_751784\|2 | ···················· | ···················· | ···················· | ···················· | ···················· | ···················· | |
| AA | 4216 | hCoV-19/USA/WV-CDC-2-3714386/2020\|EPI_ISL_751785\|2 | ···················· | ···················· | ···················· | ···················· | ···················· | ···················· | |
| AA | 4217 | hCoV-19/USA/CA-SEARCH-5389/2020\|EPI_ISL_755200\|202 | ···················· | ···················· | ···················· | ···················· | ···················· | ···················· | |
| AA | 4218 | hCoV-19/USA/CA-SEARCH-5390/2020\|EPI_ISL_755201\|202 | ···················· | ···················· | ···················· | ···················· | ···················· | ···················· | |
| AA | 4219 | hCoV-19/USA/CA-SEARCH-5391/2020\|EPI_ISL_755202\|202 | ···················· | ···················· | ···················· | ···················· | ···················· | ···················· | |
| AA | 4220 | hCoV-19/USA/CA-SEARCH-5392/2020\|EPI_ISL_755203\|202 | ···················· | ···················· | ···················· | ···················· | ···················· | ···················· | |
| AA | 4221 | hCoV-19/USA/CA-SEARCH-5393/2020\|EPI_ISL_755204\|202 | ···················· | ···················· | ···················· | ···················· | ···················· | ···················· | |
| AA | 4222 | hCoV-19/USA/CA-SEARCH-5394/2020\|EPI_ISL_755205\|202 | ···················· | ···················· | ···················· | ···················· | ···················· | ···················· | |
| AA | 4223 | hCoV-19/USA/CA-SEARCH-5395/2020\|EPI_ISL_755206\|202 | ···················· | ···················· | ···················· | ···················· | ···················· | ···················· | |
| AA | 4224 | hCoV-19/USA/CA-SEARCH-5396/2020\|EPI_ISL_755207\|202 | ···················· | ···················· | ···················· | ···················· | ···················· | ···················· | |
| AA | 4225 | hCoV-19/USA/CA-SEARCH-5397/2020\|EPI_ISL_755208\|202 | ···················· | ···················· | ···················· | ···················· | ···················· | ···················· | |
| AA | 4226 | hCoV-19/USA/CA-SEARCH-5398/2020\|EPI_ISL_755209\|202 | ············T······· | ···················· | ···················· | ···················· | ···················· | ···················· | |
| AA | 4227 | hCoV-19/Jordan/SEARCH-5399/2020\|EPI_ISL_755210\|202 | ···················· | ···················· | ···················· | ···················· | ···················· | ···················· | |
| AA | 4228 | hCoV-19/Jordan/SEARCH-5400/2020\|EPI_ISL_755211\|202 | ···················· | ···················· | ···················· | ···················· | ···················· | ···················· | |
| AA | 4229 | hCoV-19/Jordan/SEARCH-5401/2020\|EPI_ISL_755212\|202 | ···················· | ···················· | ···················· | ···················· | ···················· | ···················· | |
| AA | 4230 | hCoV-19/Jordan/SEARCH-5402/2020\|EPI_ISL_755213\|202 | ···················· | ···················· | ···················· | ···················· | ···················· | ···················· | |
| AA | 4231 | hCoV-19/Jordan/SEARCH-5403/2020\|EPI_ISL_755214\|202 | ···················· | ···················· | ···················· | ···················· | ···················· | ···················· | |
| AA | 4232 | hCoV-19/USA/CA-SEARCH-5404/2020\|EPI_ISL_755215\|202 | ···················· | ···················· | ···················· | ···················· | ···················· | ···················· | |
| AA | 4233 | hCoV-19/USA/CA-SEARCH-5405/2020\|EPI_ISL_755216\|202 | ···················· | ·A·················· | ···················· | ···················· | ···················· | ···················· | |
| AA | 4234 | hCoV-19/USA/CA-SEARCH-5406/2020\|EPI_ISL_755217\|202 | ···················· | ·A·················· | ···················· | ···················· | ···················· | ···················· | |
| AA | 4235 | hCoV-19/USA/CA-SEARCH-5407/2020\|EPI_ISL_755218\|202 | ···················· | ···················· | ···················· | ···················· | ···················· | ···················· | |
| AA | 4236 | hCoV-19/USA/CA-SEARCH-5408/2020\|EPI_ISL_755219\|202 | ···················· | ·A·················· | ···················· | ···················· | ···················· | ···················· | |
| AA | 4237 | hCoV-19/USA/CA-SEARCH-5409/2020\|EPI_ISL_755220\|202 | ···················· | ···················· | ···················· | ···················· | ···················· | ···················· | |
| AA | 4238 | hCoV-19/USA/CA-SEARCH-5410/2020\|EPI_ISL_755221\|202 | ···················· | ···················· | ···················· | ···················· | ···················· | ···················· | |
| AA | 4239 | hCoV-19/USA/CA-SEARCH-5412/2020\|EPI_ISL_755222\|202 | ···················· | ···················· | ···················· | ···················· | ···················· | ···················· | |
| AA | 4240 | hCoV-19/USA/CA-SEARCH-5413/2020\|EPI_ISL_755223\|202 | ···················· | ···················A | ···················· | ···················· | ···················· | ···················· | 1 |
| AA | 4241 | hCoV-19/USA/CA-SEARCH-5414/2020\|EPI_ISL_755224\|202 | ···················· | ···················· | ···················· | ···················· | ···················· | ···················· | |
| AA | 4242 | hCoV-19/USA/CA-SEARCH-5415/2020\|EPI_ISL_755225\|202 | ···················· | ···················· | ···················· | ···················· | ···················· | ···················· | |
| AA | 4243 | hCoV-19/USA/CA-SEARCH-5416/2020\|EPI_ISL_755226\|202 | ···················· | ···················· | ···················· | ···················· | ···················· | ···················· | |

| | | | | | | | |
|---|---|---|---|---|---|---|---|
| AA | 4244 hCoV-19/USA/CA-SEARCH-5417/2020\|EPI_ISL_755227\|202 | ···················· | ···················· | ···················· | ···················· | ···················· | ···················· |
| AA | 4245 hCoV-19/USA/CA-SEARCH-5419/2020\|EPI_ISL_755228\|202 | ···················· | ···················· | ···················· | ···················· | ···················· | ···················· |
| AA | 4246 hCoV-19/USA/CA-SEARCH-5420/2020\|EPI_ISL_755229\|202 | ···················· | ·A·················· | ···················· | ···················· | ···················· | ···················· |
| AA | 4247 hCoV-19/USA/CA-SEARCH-5421/2020\|EPI_ISL_755230\|202 | ···················· | ···················· | ···················· | ···················· | ···················· | ···················· |
| AA | 4248 hCoV-19/USA/CA-SEARCH-5422/2020\|EPI_ISL_755231\|202 | ···················· | ···················· | ···················· | ···················· | ···················· | ···················· |
| AA | 4249 hCoV-19/Jordan/SEARCH-5424/2020\|EPI_ISL_755232\|202 | ···················· | ···················· | ···················· | ···················· | ···················· | ···················· |
| AA | 4250 hCoV-19/Jordan/SEARCH-5425/2020\|EPI_ISL_755233\|202 | ···················· | ···················· | ···················· | ···················· | ···················· | ···················· |
| AA | 4251 hCoV-19/Jordan/SEARCH-5426/2020\|EPI_ISL_755234\|202 | ···················· | ···················· | ···················· | ···················· | ···················· | ···················· |
| AA | 4252 hCoV-19/Jordan/SEARCH-5427/2020\|EPI_ISL_755235\|202 | ···················· | ···················· | ···················· | ···················· | ···················· | ···················· |
| AA | 4253 hCoV-19/Jordan/SEARCH-5429/2020\|EPI_ISL_755236\|202 | ···················· | ···················· | ···················· | ···················· | ···················· | ···················· |
| AA | 4254 hCoV-19/Jordan/SEARCH-5430/2020\|EPI_ISL_755237\|202 | ···················· | ···················· | ···················· | ···················· | ···················· | ···················· |
| AA | 4255 hCoV-19/Jordan/SEARCH-5431/2020\|EPI_ISL_755238\|202 | ···················· | ···················· | ···················· | ···················· | ···················· | ···················· |
| AA | 4256 hCoV-19/Jordan/SEARCH-5432/2020\|EPI_ISL_755239\|202 | ···················· | ···················· | ···················· | ···················· | ···················· | ···················· |
| AA | 4257 hCoV-19/Jordan/SEARCH-5437/2020\|EPI_ISL_755240\|202 | ···················· | ···················· | ···················· | ···················· | ···················· | ···················· |
| AA | 4258 hCoV-19/Jordan/SEARCH-5445/2020\|EPI_ISL_755241\|202 | ···················· | ···················· | ···················· | ···················· | ···················· | ···················· |
| AA | 4259 hCoV-19/Jordan/SEARCH-5446/2020\|EPI_ISL_755242\|202 | ···················· | ···················· | ···················· | ···················· | ···················· | ···················· |
| AA | 4260 hCoV-19/Jordan/SEARCH-5451/2020\|EPI_ISL_755243\|202 | ···················· | ···················· | ···················· | ···················· | ···················· | ···················· |
| AA | 4261 hCoV-19/Jordan/SEARCH-5453/2020\|EPI_ISL_755244\|202 | ···················· | ···················· | ···················· | ···················· | ···················· | ···················· |
| AA | 4262 hCoV-19/Jordan/SEARCH-5457/2020\|EPI_ISL_755245\|202 | ···················· | ···················· | ···················· | ···················· | ···················· | ···················· |
| AA | 4263 hCoV-19/Jordan/SEARCH-5459/2020\|EPI_ISL_755246\|202 | ···················· | ···················· | ···················· | ···················· | ···················· | ···················· |
| AA | 4264 hCoV-19/Jordan/SEARCH-5460/2020\|EPI_ISL_755247\|202 | ···················· | ···················· | ···················· | ···················· | ···················· | ···················· |
| AA | 4265 hCoV-19/Jordan/SEARCH-5461/2020\|EPI_ISL_755248\|202 | ···················· | ···················· | ···················· | ···················· | ···················· | ···················· |
| AA | 4266 hCoV-19/Jordan/SEARCH-5462/2020\|EPI_ISL_755249\|202 | ···················· | ···················· | ···················· | ···················· | ···················· | ···················· |
| AA | 4267 hCoV-19/Jordan/SEARCH-5463/2020\|EPI_ISL_755250\|202 | ···················· | ···················· | ···················· | ···················· | ···················· | ···················· |
| AA | 4268 hCoV-19/Jordan/SEARCH-5465/2020\|EPI_ISL_755251\|202 | ···················· | ···················· | ···················· | ···················· | ···················· | ···················· |
| AA | 4269 hCoV-19/Jordan/SEARCH-5466/2020\|EPI_ISL_755252\|202 | ···················· | ···················· | ···················· | ···················· | ···················· | ···················· |
| AA | 4270 hCoV-19/Jordan/SEARCH-5467/2020\|EPI_ISL_755253\|202 | ···················· | ···················· | ···················· | ···················· | ···················· | ···················· |
| AA | 4271 hCoV-19/Jordan/SEARCH-5473/2020\|EPI_ISL_755254\|202 | ···················· | ···················· | ···················· | ···················· | ···················· | ···················· |
| AA | 4272 hCoV-19/Jordan/SEARCH-5474/2020\|EPI_ISL_755255\|202 | ···················· | ···················· | ···················· | ···················· | ···················· | ···················· |
| AA | 4273 hCoV-19/Jordan/SEARCH-5476/2020\|EPI_ISL_755256\|202 | ···················· | ···················· | ···················· | ···················· | ···················· | ···················· |
| AA | 4274 hCoV-19/Jordan/SEARCH-5477/2020\|EPI_ISL_755257\|202 | ···················· | ···················· | ···················· | ···················· | ···················· | ···················· |
| AA | 4275 hCoV-19/Jordan/SEARCH-5478/2020\|EPI_ISL_755258\|202 | ···················· | ···················· | ···················· | ···················· | ···················· | ···················· |
| AA | 4276 hCoV-19/Jordan/SEARCH-5479/2020\|EPI_ISL_755259\|202 | ···················· | ···················· | ···················· | ···················· | ···················· | ···················· |
| AA | 4277 hCoV-19/Jordan/SEARCH-5480/2020\|EPI_ISL_755260\|202 | ···················· | ···················· | ···················· | ···················· | ···················· | ···················· |
| AA | 4278 hCoV-19/Jordan/SEARCH-5484/2020\|EPI_ISL_755261\|202 | ···················· | ···················· | ···················· | ···················· | ···················· | ···················· |
| AA | 4279 hCoV-19/Jordan/SEARCH-5486/2020\|EPI_ISL_755262\|202 | ···················· | ···················· | ···················· | ···················· | ···················· | ···················· |
| AA | 4280 hCoV-19/Jordan/SEARCH-5490/2020\|EPI_ISL_755263\|202 | ···················· | ···················· | ···················· | ···················· | ···················· | ···················· |
| AA | 4281 hCoV-19/Jordan/SEARCH-5497/2020\|EPI_ISL_755264\|202 | ···················· | ···················· | ···················· | ···················· | ···················· | ···················· |
| AA | 4282 hCoV-19/Jordan/SEARCH-5498/2020\|EPI_ISL_755265\|202 | ···················· | ···················· | ···················· | ···················· | ···················· | ···················· |
| AA | 4283 hCoV-19/Jordan/SEARCH-5499/2020\|EPI_ISL_755266\|202 | ···················· | ···················· | ···················· | ···················· | ···················· | ···················· |
| AA | 4284 hCoV-19/Jordan/SEARCH-5500/2020\|EPI_ISL_755267\|202 | ···················· | ···················· | ···················· | ···················· | ···················· | ···················· |
| AA | 4285 hCoV-19/Jordan/SEARCH-5501/2020\|EPI_ISL_755268\|202 | ···················· | ···················· | ···················· | ···················· | ···················· | ···················· |
| AA | 4286 hCoV-19/USA/CA-SEARCH-5504/2020\|EPI_ISL_755269\|202 | ···················· | ·A·················· | ···················· | ···················· | ···················· | ···················· |
| AA | 4287 hCoV-19/USA/CA-SEARCH-5506/2020\|EPI_ISL_755270\|202 | ···················· | ···················· | ···················· | ···················· | ···················· | ···················· |
| AA | 4288 hCoV-19/USA/CA-SEARCH-5510/2020\|EPI_ISL_755271\|202 | ···················· | ···················· | ···················· | ···················· | ···················· | ···················· |
| AA | 4289 hCoV-19/USA/CA-SEARCH-5511/2020\|EPI_ISL_755272\|202 | ···················· | ·A·················· | ···················· | ···················· | ···················· | ···················· |
| AA | 4290 hCoV-19/USA/CA-SEARCH-5512/2020\|EPI_ISL_755273\|202 | ···················· | ·A·················· | ···················· | ···················· | ···················· | ···················· |
| AA | 4291 hCoV-19/USA/CA-SEARCH-5515/2020\|EPI_ISL_755274\|202 | ···················· | ·A·················· | ···················· | ···················· | ···················· | ···················· |
| AA | 4292 hCoV-19/USA/CA-SEARCH-5519/2020\|EPI_ISL_755275\|202 | ···················· | ···················· | ···················· | ···················· | ···················· | ···················· |
| AA | 4293 hCoV-19/USA/CA-SEARCH-5521/2020\|EPI_ISL_755276\|202 | ···················· | ···················· | ···················· | ···················· | ···················· | ···················· |
| AA | 4294 hCoV-19/USA/CA-SEARCH-5528/2020\|EPI_ISL_755277\|202 | ···················· | ·A·················· | ···················· | ···················· | ···················· | ···················· |
| AA | 4295 hCoV-19/USA/CA-SEARCH-5529/2020\|EPI_ISL_755278\|202 | ···················· | ···················· | ···················· | ···················· | ···················· | ···················· |
| AA | 4296 hCoV-19/USA/CA-SEARCH-5530/2020\|EPI_ISL_755279\|202 | ···················· | ·A·················· | ···················· | ···················· | ···················· | ···················· |
| AA | 4297 hCoV-19/USA/CA-SEARCH-5533/2020\|EPI_ISL_755280\|202 | ···················· | ·A·················· | ···················· | ···················· | ···················· | ···················· |
| AA | 4298 hCoV-19/USA/CA-SEARCH-5534/2020\|EPI_ISL_755281\|202 | ···················· | ···················· | ···················· | ···················· | ···················· | ···················· |
| AA | 4299 hCoV-19/USA/CA-SEARCH-5536/2020\|EPI_ISL_755282\|202 | ···················· | ···················· | ···················· | ···················· | ···················· | ···················· |
| AA | 4300 hCoV-19/USA/CA-SEARCH-5537/2020\|EPI_ISL_755283\|202 | ···················· | ·A·················· | ···················· | ···················· | ···················· | ···················· |
| AA | 4301 hCoV-19/USA/CA-SEARCH-5541/2020\|EPI_ISL_755284\|202 | ···················· | ···················· | ···················· | ···················· | ···················· | ···················· |
| AA | 4302 hCoV-19/USA/CA-SEARCH-5544/2020\|EPI_ISL_755285\|202 | ···················· | ·A·················· | ···················· | ···················· | ···················· | ···················· |
| AA | 4303 hCoV-19/USA/CA-SEARCH-5545/2020\|EPI_ISL_755286\|202 | ···················· | ·A·················· | ···················· | ···················· | ···················· | ···················· |
| AA | 4304 hCoV-19/USA/CA-SEARCH-5548/2020\|EPI_ISL_755287\|202 | ···················· | ·A·················· | ···················· | ···················· | ···················· | ···················· |
| AA | 4305 hCoV-19/USA/CA-SEARCH-5551/2020\|EPI_ISL_755288\|202 | ···················· | ·A·················· | ···················· | ···················· | ···················· | ···················· |
| AA | 4306 hCoV-19/USA/CA-SEARCH-5552/2020\|EPI_ISL_755289\|202 | ···················· | ·A·················· | ···················· | ···················· | ···················· | ···················· |
| AA | 4307 hCoV-19/USA/CA-SEARCH-5554/2020\|EPI_ISL_755290\|202 | ···················· | ·A·················· | ···················· | ···················· | ···················· | ···················· |
| AA | 4308 hCoV-19/USA/CA-SEARCH-5556/2020\|EPI_ISL_755291\|202 | ···················· | ···················· | ···················· | ···················· | ···················· | ···················· |
| AA | 4309 hCoV-19/USA/CA-SEARCH-5557/2020\|EPI_ISL_755292\|202 | ···················· | ·A·················· | ···················· | ···················· | ···················· | ···················· |
| AA | 4310 hCoV-19/USA/CA-SEARCH-5558/2020\|EPI_ISL_755293\|202 | ···················· | ···················· | ···················· | ···················· | ···················· | ···················· |
| AA | 4311 hCoV-19/USA/CA-SEARCH-5560/2020\|EPI_ISL_755294\|202 | ···················· | ·A·················· | ···················· | ···················· | ···················· | ···················· |
| AA | 4312 hCoV-19/USA/CA-SEARCH-5562/2020\|EPI_ISL_755295\|202 | ···················· | ·A·················· | ···················· | ···················· | ···················· | ···················· |
| AA | 4313 hCoV-19/USA/CA-SEARCH-5564/2020\|EPI_ISL_755296\|202 | ···················· | ·A·················· | ···················· | ···················· | ···················· | ···················· |
| AA | 4314 hCoV-19/USA/CA-SEARCH-5565/2020\|EPI_ISL_755297\|202 | ···················· | ·A·················· | ···················· | ···················· | ···················· | ···················· |
| AA | 4315 hCoV-19/USA/CA-SEARCH-5566/2020\|EPI_ISL_755298\|202 | ···················· | ·A·················· | ···················· | ···················· | ···················· | ···················· |
| AA | 4316 hCoV-19/USA/CA-SEARCH-5570/2020\|EPI_ISL_755299\|202 | ···················· | ·A·················· | ···················· | ···················· | ···················· | ···················· |
| AA | 4317 hCoV-19/England/ALDP-C8B7C0/2020\|EPI_ISL_756503\|20 | ···················· | ···················· | ···················· | ···················· | ···················· | ···················· |
| AA | 4318 hCoV-19/England/CAMC-C84F0B/2020\|EPI_ISL_756507\|20 | ···················· | ···················· | ···················· | ···················· | ···················· | ···················· |
| AA | 4319 hCoV-19/England/ALDP-C8A55E/2020\|EPI_ISL_756514\|20 | ···················· | ···················· | ···················· | ···················· | ···················· | ···················· |
| AA | 4320 hCoV-19/England/ALDP-C8B766/2020\|EPI_ISL_756520\|20 | ···················· | ···················· | ···················· | ···················· | ···················· | ···················· |
| AA | 4321 hCoV-19/England/ALDP-C8B739/2020\|EPI_ISL_756524\|20 | ···················· | ···················· | ···················· | ···················· | ···················· | ···················· |
| AA | 4322 hCoV-19/England/CAMC-C85034/2020\|EPI_ISL_756525\|20 | ···················· | ···················· | ···················· | ···················· | ···················· | ···················· |
| AA | 4323 hCoV-19/England/ALDP-C88EBF/2020\|EPI_ISL_756527\|20 | ···················· | ···················· | ···················· | ···················· | ···················· | ···················· |

AA 4324 hCoV-19/England/ALDP-C8B836/2020|EPI_ISL_756530|20 ···················· ···················· ····················· ···················· ···················· ····················
AA 4325 hCoV-19/England/ALDP-C8B65A/2020|EPI_ISL_756532|20 ···················· ···················· ····················· ···················· ···················· ····················
AA 4326 hCoV-19/England/CAMC-C84E95/2020|EPI_ISL_756535|20 ···················· ···················· ····················· ···················· ···················· ····················
AA 4327 hCoV-19/England/ALDP-C88F61/2020|EPI_ISL_756536|20 ···················· ···················· ····················· ···················· ···················· ····················
AA 4328 hCoV-19/England/ALDP-C88F8F/2020|EPI_ISL_756537|20 ···················· ···················· ····················· ···················· ···················· ····················
AA 4329 hCoV-19/England/ALDP-C8A248/2020|EPI_ISL_756539|20 ···················· ···················· ····················· ···················· ···················· ····················
AA 4330 hCoV-19/England/CAMC-C844C0/2020|EPI_ISL_756540|20 ···················· ···················· ····················· ···················· ···················· ····················
AA 4331 hCoV-19/England/ALDP-C8B60F/2020|EPI_ISL_756544|20 ···················· ···················· ····················· ···················· ···················· ····················
AA 4332 hCoV-19/England/ALDP-C8B97F/2020|EPI_ISL_756546|20 ···················· ···················· ····················· ···················· ···················· ····················
AA 4333 hCoV-19/England/CAMC-C8609D/2020|EPI_ISL_756554|20 ···················· ···················· ····················· ···················· ···················· ····················
AA 4334 hCoV-19/England/ALDP-C8B6D2/2020|EPI_ISL_756556|20 ···················· ···················· ····················· ···················· ···················· ····················
AA 4335 hCoV-19/England/ALDP-C8A57C/2020|EPI_ISL_756557|20 ···················· ···················· ····················· ···················· ···················· ····················
AA 4336 hCoV-19/England/ALDP-C8A62E/2020|EPI_ISL_756560|20 ···················· ···················· ····················· ···················· ···················· ····················
AA 4337 hCoV-19/England/ALDP-C8B6E1/2020|EPI_ISL_756563|20 ···················· ···················· ····················· ···················· ···················· ····················
AA 4338 hCoV-19/England/CAMC-C844DF/2020|EPI_ISL_756564|20 ···················· ···················· ····················· ···················· ···················· ····················
AA 4339 hCoV-19/England/ALDP-C88F34/2020|EPI_ISL_756566|20 ···················· ···················· ····················· ···················· ······T············· ····················
AA 4340 hCoV-19/England/ALDP-C8B57B/2020|EPI_ISL_756569|20 ···················· ···················· ····················· ···················· ···················· ····················
AA 4341 hCoV-19/England/CAMC-C85465/2020|EPI_ISL_756579|20 ···················· ···················· ····················· ···················· ···················· ····················
AA 4342 hCoV-19/England/ALDP-C8ABF2/2020|EPI_ISL_756583|20 ···················· ···················· ····················· ···················· ···················· ····················
AA 4343 hCoV-19/England/CAMC-C8665F/2020|EPI_ISL_756585|20 ···················· ···················· ····················· ···················· ······T············· ····················
AA 4344 hCoV-19/England/ALDP-C88F9E/2020|EPI_ISL_756587|20 ···················· ···················· ····················· ···················· ···················· ····················
AA 4345 hCoV-19/England/CAMC-C85939/2020|EPI_ISL_756589|20 ···················· ···················· ····················· ···················· ···················· ····················
AA 4346 hCoV-19/England/ALDP-C88F25/2020|EPI_ISL_756592|20 ···················· ···················· ····················· ···················· ···················· ····················
AA 4347 hCoV-19/England/CAMC-C85FEC/2020|EPI_ISL_756594|20 ···················· ···················· ····················· ···················· ···················· ····················
AA 4348 hCoV-19/England/ALDP-C8B53F/2020|EPI_ISL_756599|20 ···················· ···················· ····················· ···················· ···················· ····················
AA 4349 hCoV-19/Denmark/DCGC-21374/2020|EPI_ISL_757896|202 ···················· ···················· ····················· ···················· ···················· ····················
AA 4350 hCoV-19/Denmark/DCGC-21382/2020|EPI_ISL_757897|202 ···················· ···················· ····················· ···················· ···················· ····················
AA 4351 hCoV-19/Denmark/DCGC-21422/2020|EPI_ISL_757898|202 ···················· ···················· ····················· ···················· ···················· ····················
AA 4352 hCoV-19/Denmark/DCGC-21429/2020|EPI_ISL_757899|202 ···················· ···················· ····················· ···················· ···················· ····················
AA 4353 hCoV-19/Denmark/DCGC-20317/2020|EPI_ISL_748110|202 ···················· ···················· ····················· ···················· ···················· ····················
AA 4354 hCoV-19/Denmark/DCGC-20318/2020|EPI_ISL_748111|202 ···················· ···················· ····················· ···················· ···················· ····················
AA 4355 hCoV-19/Denmark/DCGC-20319/2020|EPI_ISL_748112|202 ···················· ···················· ····················· ···················· ···················· ····················
AA 4356 hCoV-19/Denmark/DCGC-20320/2020|EPI_ISL_748113|202 ···················· ···················· ····················· ···················· ···················· ····················
AA 4357 hCoV-19/Denmark/DCGC-20324/2020|EPI_ISL_748114|202 ···················· ···················· ····················· ···················· ···················· ····················
AA 4358 hCoV-19/Denmark/DCGC-20326/2020|EPI_ISL_748115|202 ···················· ···················· ····················· ···················· ···················· ····················
AA 4359 hCoV-19/Denmark/DCGC-20327/2020|EPI_ISL_748116|202 ···················· ···················· ····················· ···················· ···················· ····················
AA 4360 hCoV-19/Denmark/DCGC-20332/2020|EPI_ISL_748117|202 ···················· ···················· ····················· ···················· ···················· ····················
AA 4361 hCoV-19/Denmark/DCGC-20333/2020|EPI_ISL_748118|202 ···················· ···················· ····················· ···················· ···················· ····················
AA 4362 hCoV-19/Denmark/DCGC-20335/2020|EPI_ISL_748119|202 ···················· ···················· ····················· ···················· ···················· ····················
AA 4363 hCoV-19/Denmark/DCGC-20337/2020|EPI_ISL_748120|202 ···················· ···················· ····················· ···················· ···················· ····················
AA 4364 hCoV-19/Denmark/DCGC-20338/2020|EPI_ISL_748121|202 ···················· ·A·················· ····················· ···················· ···················· ····················
AA 4365 hCoV-19/Denmark/DCGC-20339/2020|EPI_ISL_748122|202 ···················· ···················· ····················· ···················· ···················· ····················
AA 4366 hCoV-19/Denmark/DCGC-20340/2020|EPI_ISL_748123|202 ···················· ···················· ····················· ···················· ···················· ····················
AA 4367 hCoV-19/Denmark/DCGC-20343/2020|EPI_ISL_748124|202 ···················· ·········A·········· ····················· ···················· ···················· ····················
AA 4368 hCoV-19/Denmark/DCGC-20344/2020|EPI_ISL_748125|202 ···················· ···················· ····················· ···················· ···················· ····················
AA 4369 hCoV-19/Denmark/DCGC-20345/2020|EPI_ISL_748126|202 ···················· ···················· ····················· ···················· ···················· ····················
AA 4370 hCoV-19/Denmark/DCGC-20346/2020|EPI_ISL_748127|202 ···················· ···················· ····················· ···················· ···················· ····················
AA 4371 hCoV-19/Denmark/DCGC-20347/2020|EPI_ISL_748128|202 ···················· ···················· ····················· ···················· ···················· ····················
AA 4372 hCoV-19/Denmark/DCGC-20348/2020|EPI_ISL_748129|202 ···················· ···················· ····················· ···················· ···················· ····················
AA 4373 hCoV-19/Denmark/DCGC-20349/2020|EPI_ISL_748130|202 ···················· ···················· ····················· ···················· ···················· ····················
AA 4374 hCoV-19/Denmark/DCGC-20351/2020|EPI_ISL_748131|202 ···················· ···················· ····················· ···················· ···················· ····················
AA 4375 hCoV-19/Denmark/DCGC-20352/2020|EPI_ISL_748132|202 ···················· ···················· ····················· ···················· ···················· ····················
AA 4376 hCoV-19/Denmark/DCGC-20353/2020|EPI_ISL_748133|202 ···················· ···················· ····················· ···················· ···················· ····················
AA 4377 hCoV-19/Denmark/DCGC-20354/2020|EPI_ISL_748134|202 ···················· ···················· ····················· ···················· ···················· ····················
AA 4378 hCoV-19/Denmark/DCGC-20355/2020|EPI_ISL_748135|202 ···················· ···················· ····················· ···················· ···················· ····················
AA 4379 hCoV-19/Denmark/DCGC-20357/2020|EPI_ISL_748136|202 ···················· ···················· ····················· ···················· ···················· ····················
AA 4380 hCoV-19/Denmark/DCGC-20358/2020|EPI_ISL_748137|202 ···················· ···················· ····················· ···················· ···················· ····················
AA 4381 hCoV-19/Uruguay/RIV-M8/2020|EPI_ISL_748138|2020-05 ···················· ···················· ····················· ···················· ···················· ····················
AA 4382 hCoV-19/Uruguay/RIV-M9/2020|EPI_ISL_748139|2020-05 ···················· ···················· ····················· ···················· ···················· ····················
AA 4383 hCoV-19/Uruguay/RIV-M10/2020|EPI_ISL_748140|2020-0 ···················· ···················· ····················· ···················· ···················· ····················
AA 4384 hCoV-19/Uruguay/RIV-M11/2020|EPI_ISL_748141|2020-0 ···················· ···················· ····················· ···················· ···················· ····················
AA 4385 hCoV-19/Uruguay/RIV-M12/2020|EPI_ISL_748142|2020-0 ···················· ···················· ····················· ···················· ···················· ····················
AA 4386 hCoV-19/Uruguay/RIV-M13/2020|EPI_ISL_748143|2020-0 ···················· ···················· ····················· ···················· ···················· ····················
AA 4387 hCoV-19/Uruguay/RIV-M14/2020|EPI_ISL_748144|2020-0 ···················· ···················· ····················· ···················· ···················· ····················
AA 4388 hCoV-19/Uruguay/RIV-M15/2020|EPI_ISL_748145|2020-0 ···················· ···················· ····················· ···················· ···················· ····················
AA 4389 hCoV-19/Denmark/DCGC-18492/2020|EPI_ISL_749400|202 ···················· ···················· ····················· ···················· ···················· ····················
AA 4390 hCoV-19/Denmark/DCGC-18507/2020|EPI_ISL_749401|202 ···················· ···················· ····················· ···················· ···················· ····················
AA 4391 hCoV-19/Denmark/DCGC-18589/2020|EPI_ISL_749402|202 ···················· ···················· ····················· ···················· ···················· ····················
AA 4392 hCoV-19/Denmark/DCGC-18672/2020|EPI_ISL_749403|202 ···················· ···················· ····················· ···················· ···················· ····················
AA 4393 hCoV-19/Denmark/DCGC-18673/2020|EPI_ISL_749404|202 ···················· ···················· ····················· ···················· ···················· ····················
AA 4394 hCoV-19/Denmark/DCGC-18691/2020|EPI_ISL_749405|202 ···················· ···················· ····················· ···················· ···················· ····················
AA 4395 hCoV-19/Denmark/DCGC-18703/2020|EPI_ISL_749406|202 ···················· ···················· ····················· ···················· ···················· ····················
AA 4396 hCoV-19/Denmark/DCGC-17750/2020|EPI_ISL_749407|202 ···················· ···················· ····················· ···················· ···················· ····················
AA 4397 hCoV-19/Denmark/DCGC-17766/2020|EPI_ISL_749408|202 ···················· ···················· ····················· ···················· ···················· ····················
AA 4398 hCoV-19/Denmark/DCGC-17799/2020|EPI_ISL_749409|202 ···················· ···················· ····················· ···················· ···················· ····················
AA 4399 hCoV-19/Denmark/DCGC-18130/2020|EPI_ISL_749440|202 ···················· ···················· ····················· ···················· ···················· ····················
AA 4400 hCoV-19/Denmark/DCGC-18135/2020|EPI_ISL_749441|202 ···················· ···················· ····················· ···················· ···················· ····················
AA 4401 hCoV-19/Denmark/DCGC-18138/2020|EPI_ISL_749442|202 ···················· ···················· ····················· ···················· ···················· ····················
AA 4402 hCoV-19/Denmark/DCGC-18159/2020|EPI_ISL_749443|202 ···················· ···················· ····················· ···················· ···················· ····················
AA 4403 hCoV-19/Denmark/DCGC-18178/2020|EPI_ISL_749444|202 ···················· ···················· ····················· ···················· ···C················ ····················

| | | | | | | | |
|---|---|---|---|---|---|---|---|
| AA | 4404 hCoV-19/Denmark/DCGC-18183/2020\|EPI_ISL_749445\|202 | ···················· | ···················· | ···················· | ···················· | ···················· | ···················· |
| AA | 4405 hCoV-19/Denmark/DCGC-18191/2020\|EPI_ISL_749446\|202 | ···················· | ···················· | ···················· | ···················· | ···················· | ···················· |
| AA | 4406 hCoV-19/Denmark/DCGC-18204/2020\|EPI_ISL_749447\|202 | ···················· | ···················· | ···················· | ···················· | ···················· | ···················· |
| AA | 4407 hCoV-19/Denmark/DCGC-18243/2020\|EPI_ISL_749448\|202 | ···················· | ···················· | ···················· | ···················· | ···················· | ···················· |
| AA | 4408 hCoV-19/Denmark/DCGC-18256/2020\|EPI_ISL_749449\|202 | ···················· | ···················· | ···················· | ···················· | ···················· | ···················· |
| AA | 4409 hCoV-19/Denmark/DCGC-18275/2020\|EPI_ISL_749450\|202 | ···················· | ···················· | ···················· | ···················· | ···················· | ···················· |
| AA | 4410 hCoV-19/Denmark/DCGC-18279/2020\|EPI_ISL_749451\|202 | ···················· | ···················· | ···················· | ···················· | ···················· | ···················· |
| AA | 4411 hCoV-19/Denmark/DCGC-18286/2020\|EPI_ISL_749452\|202 | ···················· | ···················· | ···················· | ···················· | ···················· | ···················· |
| AA | 4412 hCoV-19/Denmark/DCGC-18287/2020\|EPI_ISL_749453\|202 | ···················· | ···················· | ···················· | ···················· | ···················· | ···················· |
| AA | 4413 hCoV-19/Denmark/DCGC-18312/2020\|EPI_ISL_749454\|202 | ···················· | ···················· | ···················· | ···················· | ···················· | ···················· |
| AA | 4414 hCoV-19/Denmark/DCGC-18319/2020\|EPI_ISL_749455\|202 | ···················· | ···················· | ···················· | ···················· | ···················· | ···················· |
| AA | 4415 hCoV-19/Denmark/DCGC-18373/2020\|EPI_ISL_749456\|202 | ···················· | ···················· | ···················· | ···················· | ···················· | ···················· |
| AA | 4416 hCoV-19/Denmark/DCGC-18377/2020\|EPI_ISL_749457\|202 | ···················· | ···················· | ···················· | ···················· | ···················· | ···················· |
| AA | 4417 hCoV-19/Denmark/DCGC-18389/2020\|EPI_ISL_749458\|202 | ···················· | ···················· | ···················· | ···················· | ···················· | ···················· |
| AA | 4418 hCoV-19/Denmark/DCGC-18411/2020\|EPI_ISL_749459\|202 | ···················· | ···················· | ···················· | ···················· | ···················· | ···················· |
| AA | 4419 hCoV-19/Denmark/DCGC-18436/2020\|EPI_ISL_749460\|202 | ···················· | ···················· | ···················· | ···················· | ···················· | ···················· |
| AA | 4420 hCoV-19/Denmark/DCGC-18441/2020\|EPI_ISL_749461\|202 | ···················· | ···················· | ···················· | ···················· | ···················· | ···················· |
| AA | 4421 hCoV-19/Denmark/DCGC-18445/2020\|EPI_ISL_749462\|202 | ···················· | ···················· | ···················· | ···················· | ···················· | ···················· |
| AA | 4422 hCoV-19/Denmark/DCGC-18464/2020\|EPI_ISL_749463\|202 | ···················· | ···················· | ···················· | ···················· | ···················· | ···················· |
| AA | 4423 hCoV-19/Denmark/DCGC-18511/2020\|EPI_ISL_749464\|202 | ···················· | ···················· | ···················· | ···················· | ···················· | ···················· |
| AA | 4424 hCoV-19/Denmark/DCGC-18533/2020\|EPI_ISL_749465\|202 | ···················· | ···················· | ···················· | ···················· | ···················· | ···················· |
| AA | 4425 hCoV-19/Denmark/DCGC-18547/2020\|EPI_ISL_749466\|202 | ···················· | ···················· | ···················· | ···················· | ···················· | ···················· |
| AA | 4426 hCoV-19/Denmark/DCGC-18572/2020\|EPI_ISL_749467\|202 | ···················· | ···················· | ···················· | ···················· | ···················· | ···················· |
| AA | 4427 hCoV-19/Denmark/DCGC-18619/2020\|EPI_ISL_749468\|202 | ···················· | ···················· | ···················· | ···················· | ···················· | ···················· |
| AA | 4428 hCoV-19/Denmark/DCGC-18639/2020\|EPI_ISL_749469\|202 | ···················· | ···················· | ···················· | ···················· | ···················· | ···················· |
| AA | 4429 hCoV-19/Denmark/DCGC-18641/2020\|EPI_ISL_749470\|202 | ···················· | ···················· | ···················· | ···················· | ···················· | ···················· |
| AA | 4430 hCoV-19/Denmark/DCGC-18650/2020\|EPI_ISL_749471\|202 | ···················· | ···················· | ···················· | ···················· | ···················· | ···················· |
| AA | 4431 hCoV-19/Denmark/DCGC-18652/2020\|EPI_ISL_749472\|202 | ···················· | ···················· | ···················· | ···················· | ··C················· | ···················· |
| AA | 4432 hCoV-19/Denmark/DCGC-18658/2020\|EPI_ISL_749473\|202 | ···················· | ···················· | ···················· | ···················· | ···················· | ···················· |
| AA | 4433 hCoV-19/Denmark/DCGC-18668/2020\|EPI_ISL_749475\|202 | ···················· | ···················· | ···················· | ···················· | ···················· | ···················· |
| AA | 4434 hCoV-19/Denmark/DCGC-18679/2020\|EPI_ISL_749476\|202 | ···················· | ···················· | ···················· | ···················· | ···················· | ···················· |
| AA | 4435 hCoV-19/Denmark/DCGC-18683/2020\|EPI_ISL_749477\|202 | ···················· | ···················· | ···················· | ···················· | ···················· | ···················· |
| AA | 4436 hCoV-19/Denmark/DCGC-18684/2020\|EPI_ISL_749478\|202 | ···················· | ···················· | ···················· | ···················· | ···················· | ···················· |
| AA | 4437 hCoV-19/Denmark/DCGC-18715/2020\|EPI_ISL_749479\|202 | ···················· | ···················· | ···················· | ···················· | ···················· | ···················· |
| AA | 4438 hCoV-19/Denmark/DCGC-17115/2020\|EPI_ISL_750400\|202 | ···················· | ···················· | ···················· | ···················· | ···················· | ···················· |
| AA | 4439 hCoV-19/Denmark/DCGC-17353/2020\|EPI_ISL_750401\|202 | ···················· | ···················· | ···················· | ···················· | ···················· | ···················· |
| AA | 4440 hCoV-19/Denmark/DCGC-17420/2020\|EPI_ISL_750402\|202 | ···················· | ···················· | ···················· | ···················· | ···················· | ···················· |
| AA | 4441 hCoV-19/Denmark/DCGC-17450/2020\|EPI_ISL_750403\|202 | ···················· | ···················· | ···················· | ···················· | ···················· | ···················· |
| AA | 4442 hCoV-19/Denmark/DCGC-17662/2020\|EPI_ISL_750404\|202 | ···················· | ···················· | ···················· | ···················· | ···················· | ···················· |
| AA | 4443 hCoV-19/Denmark/DCGC-17698/2020\|EPI_ISL_750405\|202 | ···················· | ···················· | ···················· | ···················· | ···················· | ···················· |
| AA | 4444 hCoV-19/Denmark/DCGC-17714/2020\|EPI_ISL_750406\|202 | ···················· | ···················· | ···················· | ···················· | ···················· | ···················· |
| AA | 4445 hCoV-19/Denmark/DCGC-16760/2020\|EPI_ISL_750407\|202 | ···················· | ···················· | ···················· | ···················· | ···················· | ···················· |
| AA | 4446 hCoV-19/Denmark/DCGC-16764/2020\|EPI_ISL_750408\|202 | ···················· | ···················· | ···················· | ···················· | ···················· | ···················· |
| AA | 4447 hCoV-19/Denmark/DCGC-16769/2020\|EPI_ISL_750409\|202 | ···················· | ···················· | ···················· | ···················· | ···················· | ···················· |
| AA | 4448 hCoV-19/Denmark/DCGC-16805/2020\|EPI_ISL_750410\|202 | ···················· | ···················· | ···················· | ···················· | ···················· | ···················· |
| AA | 4449 hCoV-19/Denmark/DCGC-16806/2020\|EPI_ISL_750411\|202 | ···················· | ···················· | ···················· | ···················· | ···················· | ···················· |
| AA | 4450 hCoV-19/Denmark/DCGC-16831/2020\|EPI_ISL_750412\|202 | ···················· | ···················· | ···················· | ···················· | ···················· | ···················· |
| AA | 4451 hCoV-19/Denmark/DCGC-16855/2020\|EPI_ISL_750413\|202 | ···················· | ···················· | ···················· | ···················· | ···················· | ···················· |
| AA | 4452 hCoV-19/Denmark/DCGC-16864/2020\|EPI_ISL_750414\|202 | ···················· | ···················· | ···················· | ···················· | ···················· | ···················· |
| AA | 4453 hCoV-19/Denmark/DCGC-16867/2020\|EPI_ISL_750415\|202 | ···················· | ···················· | ···················· | ···················· | ···················· | ···················· |
| AA | 4454 hCoV-19/Denmark/DCGC-16870/2020\|EPI_ISL_750416\|202 | ···················· | ···················· | ···················· | ···················· | ···················· | ···················· |
| AA | 4455 hCoV-19/Denmark/DCGC-16880/2020\|EPI_ISL_750417\|202 | ···················· | ···················· | ···················· | ···················· | ···················· | ···················· |
| AA | 4456 hCoV-19/Denmark/DCGC-16918/2020\|EPI_ISL_750418\|202 | ···················· | ···················· | ···················· | ···················· | ···················· | ···················· |
| AA | 4457 hCoV-19/Denmark/DCGC-16926/2020\|EPI_ISL_750419\|202 | ···················· | ···················· | ···················· | ···················· | ···················· | ···················· |
| AA | 4458 hCoV-19/Denmark/DCGC-16937/2020\|EPI_ISL_750420\|202 | ···················· | ···················· | ···················· | ···················· | ···················· | ···················· |
| AA | 4459 hCoV-19/Denmark/DCGC-16942/2020\|EPI_ISL_750421\|202 | ···················· | ···················· | ···················· | ···················· | ···················· | ···················· |
| AA | 4460 hCoV-19/Denmark/DCGC-17294/2020\|EPI_ISL_750450\|202 | ···················· | ···················· | ···················· | ···················· | ···················· | ···················· |
| AA | 4461 hCoV-19/Denmark/DCGC-17315/2020\|EPI_ISL_750452\|202 | ···················· | ···················· | ···················· | ···················· | ···················· | ···················· |
| AA | 4462 hCoV-19/Denmark/DCGC-17346/2020\|EPI_ISL_750455\|202 | ···················· | ···················· | ···················· | ···················· | ···················· | ···················· |
| AA | 4463 hCoV-19/Denmark/DCGC-17355/2020\|EPI_ISL_750456\|202 | ···················· | ···················· | ···················· | ···················· | ···················· | ···················· |
| AA | 4464 hCoV-19/Denmark/DCGC-17368/2020\|EPI_ISL_750457\|202 | ···················· | ···················· | ···················· | ···················· | ···················· | ···················· |
| AA | 4465 hCoV-19/Denmark/DCGC-17370/2020\|EPI_ISL_750458\|202 | ···················· | ···················· | ···················· | ···················· | ···················· | ···················· |
| AA | 4466 hCoV-19/Denmark/DCGC-17380/2020\|EPI_ISL_750459\|202 | ···················· | ···················· | ···················· | ···················· | ···················· | ···················· |
| AA | 4467 hCoV-19/Denmark/DCGC-17397/2020\|EPI_ISL_750460\|202 | ···················· | ···················· | ···················· | ···················· | ···················· | ···················· |
| AA | 4468 hCoV-19/Denmark/DCGC-17411/2020\|EPI_ISL_750461\|202 | ···················· | ···················· | ···················· | ···················· | ···················· | ···················· |
| AA | 4469 hCoV-19/Denmark/DCGC-17423/2020\|EPI_ISL_750462\|202 | ···················· | ···················· | ···················· | ···················· | ···················· | ···················· |
| AA | 4470 hCoV-19/Denmark/DCGC-17453/2020\|EPI_ISL_750463\|202 | ···················· | ···················· | ···················· | ···················· | ···················· | ···················· |
| AA | 4471 hCoV-19/Denmark/DCGC-17455/2020\|EPI_ISL_750464\|202 | ···················· | ···················· | ···················· | ···················· | ···················· | ···················· |
| AA | 4472 hCoV-19/Denmark/DCGC-17458/2020\|EPI_ISL_750465\|202 | ···················· | ···················· | ···················· | ···················· | ···················· | ···················· |
| AA | 4473 hCoV-19/USA/MT-CDC-2-3714176/2020\|EPI_ISL_751710\|2 | ···················· | ···················· | ···················· | ···················· | ···················· | ···················· |
| AA | 4474 hCoV-19/USA/MT-CDC-2-3714181/2020\|EPI_ISL_751711\|2 | ···················· | ···················· | ···················· | ···················· | ···················· | ···················· |
| AA | 4475 hCoV-19/USA/NH-CDC-2-3714193/2020\|EPI_ISL_751712\|2 | ···················· | ···················· | ···················· | ···················· | ···················· | ···················· |
| AA | 4476 hCoV-19/USA/MT-CDC-2-3714194/2020\|EPI_ISL_751713\|2 | ···················· | ···················· | ···················· | ···················· | ···················· | ···················· |
| AA | 4477 hCoV-19/USA/NV-CDC-2-3714253/2020\|EPI_ISL_751714\|2 | ···················· | ·A·················· | ···················· | ···················· | ···················· | ···················· |
| AA | 4478 hCoV-19/USA/MO-CDC-2-3714256/2020\|EPI_ISL_751715\|2 | ········T··········· | ·A·················· | ···················· | ···················· | ···················· | ···················· |
| AA | 4479 hCoV-19/USA/MN-CDC-2-3714263/2020\|EPI_ISL_751716\|2 | ···················· | ···················· | ···················· | ···················· | ···················· | ···················· |
| AA | 4480 hCoV-19/USA/NY-CDC-2-3714264/2020\|EPI_ISL_751717\|2 | ···················· | ···················· | ···················· | ···················· | ···················· | ···················· |
| AA | 4481 hCoV-19/USA/SC-CDC-2-3714351/2020\|EPI_ISL_751718\|2 | ···················· | ·A·················· | ···················· | ···················· | ···················· | ···················· |
| AA | 4482 hCoV-19/USA/ID-CDC-2-3714381/2020\|EPI_ISL_751719\|2 | ···················· | ···················· | ···················· | ···················· | ···················· | ···················· |
| AA | 4483 hCoV-19/USA/TX-CDC-2-3714385/2020\|EPI_ISL_751720\|2 | ···················· | ···················· | ···················· | ···················· | ···················· | ···················· |

| | | | | | | | |
|---|---|---|---|---|---|---|---|
| AA | 4484 hCoV-19/USA/IA-CDC-2-3714458/2020\|EPI_ISL_751721\|2 | ···················· | ···················· | ····················· | ···················· | ···················· | ···················· |
| AA | 4485 hCoV-19/USA/LA-CDC-2-3714570/2020\|EPI_ISL_751722\|2 | ···················· | ·A·················· | ····················· | ···················· | ···················· | ···················· |
| AA | 4486 hCoV-19/USA/AR-CDC-2-3713955/2020\|EPI_ISL_751723\|2 | ···················· | ···················· | ····················· | ···················· | ···················· | ···················· |
| AA | 4487 hCoV-19/USA/KS-CDC-2-3713958/2020\|EPI_ISL_751724\|2 | ···················· | ···················· | ····················· | ···················· | ···················· | ···················· |
| AA | 4488 hCoV-19/USA/MD-CDC-2-3713969/2020\|EPI_ISL_751725\|2 | ···················· | ···················· | ····················· | ···················· | ···················· | ···················· |
| AA | 4489 hCoV-19/USA/DE-CDC-2-3713996/2020\|EPI_ISL_751726\|2 | ···················· | ···················· | ····················· | ···················· | ···················· | ···················· |
| AA | 4490 hCoV-19/USA/IN-CDC-2-3714056/2020\|EPI_ISL_751727\|2 | ···················· | ··C················· | ····················· | ···················· | ···················· | ···················· |
| AA | 4491 hCoV-19/USA/CO-CDC-2-3714069/2020\|EPI_ISL_751728\|2 | ···················· | ···················· | ····················· | ···················· | ···················· | ···················· |
| AA | 4492 hCoV-19/USA/CT-CDC-2-3714070/2020\|EPI_ISL_751729\|2 | ···················· | ···················· | ····················· | ···················· | ···················· | ···················· |
| AA | 4493 hCoV-19/USA/KS-CDC-2-3714073/2020\|EPI_ISL_751730\|2 | ···················· | ···················· | ····················· | ···················· | ···················· | ···················· |
| AA | 4494 hCoV-19/USA/AR-CDC-2-3714076/2020\|EPI_ISL_751731\|2 | ···················· | ···················· | ····················· | ···················· | ···················· | ···················· |
| AA | 4495 hCoV-19/USA/NE-CDC-2-3714172/2020\|EPI_ISL_751732\|2 | ···················· | ···················· | ····················· | ···················· | ···················· | ···················· |
| AA | 4496 hCoV-19/USA/MN-CDC-2-3714173/2020\|EPI_ISL_751733\|2 | ···················· | ···················· | ····················· | ···················· | ···················· | ···················· |
| AA | 4497 hCoV-19/USA/NE-CDC-2-3714177/2020\|EPI_ISL_751734\|2 | ···················· | ···················· | ····················· | ···················· | ···················· | ···················· |
| AA | 4498 hCoV-19/USA/NH-CDC-2-3714182/2020\|EPI_ISL_751735\|2 | ···················· | ·············A······ | ····················· | ···················· | ···················· | ···················· |
| AA | 4499 hCoV-19/USA/NV-CDC-2-3714189/2020\|EPI_ISL_751736\|2 | ···················· | ···················· | ····················· | ···················· | ···················· | ···················· |
| AA | 4500 hCoV-19/USA/MO-CDC-2-3714191/2020\|EPI_ISL_751737\|2 | ···················· | ···················· | ····················· | ···················· | ···················· | ···················· |
| AA | 4501 hCoV-19/USA/MT-CDC-2-3714259/2020\|EPI_ISL_751738\|2 | ···················· | ···················· | ····················· | ···················· | ···················· | ···················· |
| AA | 4502 hCoV-19/USA/MN-CDC-2-3714271/2020\|EPI_ISL_751739\|2 | ···················· | ···················· | ····················· | ···················· | ···················· | ···················· |
| AA | 4503 hCoV-19/USA/MO-CDC-2-3714287/2020\|EPI_ISL_751740\|2 | ···················· | ···················· | ····················· | ···················· | ···················· | ···················· |
| AA | 4504 hCoV-19/USA/OH-CDC-2-3714358/2020\|EPI_ISL_751741\|2 | ···················· | ···················· | ····················· | ···················· | ···················· | ···················· |
| AA | 4505 hCoV-19/USA/IA-CDC-2-3714359/2020\|EPI_ISL_751742\|2 | ···················· | ···················· | ····················· | ···················· | ···················· | ···················· |
| AA | 4506 hCoV-19/USA/WY-CDC-2-3714364/2020\|EPI_ISL_751743\|2 | ···················· | ···················· | ····················· | ···················· | ···················· | ···················· |
| AA | 4507 hCoV-19/USA/SC-CDC-2-3714370/2020\|EPI_ISL_751744\|2 | ···················· | ·A·················· | ····················· | ···················· | ···················· | ···················· |
| AA | 4508 hCoV-19/USA/VA-CDC-2-3714372/2020\|EPI_ISL_751745\|2 | ···················· | ···················· | ····················· | ···················· | ···················· | ···················· |
| AA | 4509 hCoV-19/USA/WV-CDC-2-3714397/2020\|EPI_ISL_751746\|2 | ···················· | ···················· | ····················· | ···················· | ···················· | ···················· |
| AA | 4510 hCoV-19/USA/VA-CDC-2-3714461/2020\|EPI_ISL_751747\|2 | ···················· | ·A·················· | ····················· | ···················· | ···················· | ···················· |
| AA | 4511 hCoV-19/USA/ID-CDC-2-3714462/2020\|EPI_ISL_751748\|2 | ···················· | ···················· | ····················· | ···················· | ···················· | ···················· |
| AA | 4512 hCoV-19/USA/TX-CDC-2-3714479/2020\|EPI_ISL_751749\|2 | ···················· | ···················· | ····················· | ···················· | ···················· | ···················· |
| AA | 4513 hCoV-19/USA/ID-CDC-2-3714480/2020\|EPI_ISL_751750\|2 | ···················· | ···················· | ····················· | ···················· | ···················· | ···················· |
| AA | 4514 hCoV-19/USA/RI-CDC-2-3714487/2020\|EPI_ISL_751751\|2 | ···················· | ···················· | ····················· | ···················· | ···················· | ···················· |
| AA | 4515 hCoV-19/USA/LA-CDC-2-3714499/2020\|EPI_ISL_751752\|2 | ···················· | ···················· | ····················· | ···················· | ···················· | ···················· |
| AA | 4516 hCoV-19/USA/ID-CDC-2-3714353/2020\|EPI_ISL_751780\|2 | ···················· | ···················· | ····················· | ···················· | ···················· | ···················· |
| AA | 4517 hCoV-19/USA/WY-CDC-2-3714357/2020\|EPI_ISL_751781\|2 | ···················· | ······A············· | ····················· | ···················· | ···················· | ···················· |
| AA | 4518 hCoV-19/USA/WV-CDC-2-3714390/2020\|EPI_ISL_751786\|2 | ···················· | ···················· | ····················· | ···················· | ···················· | ···················· |
| AA | 4519 hCoV-19/USA/TX-CDC-2-3714457/2020\|EPI_ISL_751787\|2 | ···················· | ···················· | ····················· | ···················· | ···················· | ···················· |
| AA | 4520 hCoV-19/USA/OH-CDC-2-3714473/2020\|EPI_ISL_751788\|2 | ···················· | ·············A······ | ····················· | ···················· | ···················· | ···················· |
| AA | 4521 hCoV-19/USA/ID-CDC-2-3714481/2020\|EPI_ISL_751789\|2 | ···················· | ···················· | ····················· | ···················· | ···················· | ···················· |
| AA | 4522 hCoV-19/USA/RI-CDC-2-3714490/2020\|EPI_ISL_751790\|2 | ···················· | ···················· | ····················· | ···················· | ···················· | ···················· |
| AA | 4523 hCoV-19/USA/AL-CDC-2-3714495/2020\|EPI_ISL_751791\|2 | ···················· | ·A·················· | ····················· | ···················· | ···················· | ···················· |
| AA | 4524 hCoV-19/USA/AL-CDC-2-3714557/2020\|EPI_ISL_751792\|2 | ···················· | ···················· | ····················· | ···················· | ···················· | ···················· |
| AA | 4525 hCoV-19/USA/AK-CDC-2-3714567/2020\|EPI_ISL_751793\|2 | ···················· | ···················· | ····················· | ···················· | ···················· | ···················· |
| AA | 4526 hCoV-19/USA/KY-CDC-2-3714568/2020\|EPI_ISL_751794\|2 | ···················· | ·A·················· | ····················· | ···················· | ···················· | ···················· |
| AA | 4527 hCoV-19/USA/RI-CDC-2-3714578/2020\|EPI_ISL_751795\|2 | ···················· | ···················· | ····················· | ···················· | ···················· | ···················· |
| AA | 4528 hCoV-19/USA/CA-CHLA-PLM71343358/2020\|EPI_ISL_75353 | ···················· | ···················· | ····················· | ···················· | ···················· | ···················· |
| AA | 4529 hCoV-19/USA/CA-CHLA-PLM54586330/2020\|EPI_ISL_75353 | ···················· | ···················· | ····················· | ···················· | ···················· | ···················· |
| AA | 4530 hCoV-19/USA/CA-CHLA-PLM07635073/2020\|EPI_ISL_75353 | ···················· | ···················· | ····················· | ···················· | ···················· | ···················· |
| AA | 4531 hCoV-19/USA/CA-CHLA-PLM12515819/2020\|EPI_ISL_75353 | ···················· | ···················· | ····················· | ···················· | ···················· | ···················· |
| AA | 4532 hCoV-19/USA/CA-CHLA-PLM73511071/2020\|EPI_ISL_75353 | ···················· | ····A··············· | ····················· | ···················· | ···················· | ···················· |
| AA | 4533 hCoV-19/USA/CA-CHLA-PLM57647613/2020\|EPI_ISL_75354 | ···················· | ···················· | ····················· | ···················· | ···················· | ···················· |
| AA | 4534 hCoV-19/USA/CA-CHLA-PLM73838575/2020\|EPI_ISL_75354 | ···················· | ···················· | ····················· | ···················· | ···················· | ···················· |
| AA | 4535 hCoV-19/USA/CA-CHLA-PLM78910262/2020\|EPI_ISL_75354 | ···················· | ···················· | ····················· | ···················· | ···················· | ···················· |
| AA | 4536 hCoV-19/USA/CA-CHLA-PLM50751246/2020\|EPI_ISL_75354 | ···················· | ···················· | ····················· | ···················· | ···················· | ···················· |
| AA | 4537 hCoV-19/USA/CA-CHLA-PLM07223004/2020\|EPI_ISL_75354 | ···················· | ···················· | ····················· | ···················· | ···················· | ···················· |
| AA | 4538 hCoV-19/USA/CA-CHLA-PLM62737694/2020\|EPI_ISL_75354 | ···················· | ···················· | ····················· | ···················· | ···················· | ···················· |
| AA | 4539 hCoV-19/USA/CA-CHLA-PLM65980412/2020\|EPI_ISL_75354 | ···················· | ···················· | ····················· | ···················· | ···················· | ···················· |
| AA | 4540 hCoV-19/USA/CA-CHLA-PLM05944876/2020\|EPI_ISL_75354 | ···················· | ·A·················· | ····················· | ···················· | ···················· | ···················· |
| AA | 4541 hCoV-19/USA/CA-CHLA-PLM49510562/2020\|EPI_ISL_75354 | ···················· | ·A·················· | ····················· | ···················· | ···················· | ···················· |
| AA | 4542 hCoV-19/USA/CA-CHLA-PLM77346599/2020\|EPI_ISL_75354 | ···················· | ···················· | ····················· | ···················· | ···················· | ···················· |
| AA | 4543 hCoV-19/USA/CA-CHLA-PLM76481572/2020\|EPI_ISL_75355 | ···················· | ···················· | ····················· | ···················· | ···················· | ···················· |
| AA | 4544 hCoV-19/USA/CA-CHLA-PLM92463629/2020\|EPI_ISL_75355 | ···················· | ·A·················· | ····················· | ···················· | ···················· | ···················· |
| AA | 4545 hCoV-19/USA/CA-CHLA-PLM78606325/2020\|EPI_ISL_75355 | ···················· | ···················· | ····················· | ···················· | ···················· | ···················· |
| AA | 4546 hCoV-19/USA/CA-CHLA-PLM02423366/2020\|EPI_ISL_75355 | ···················· | ···················· | ····················· | ···················· | ···················· | ···················· |
| AA | 4547 hCoV-19/USA/CA-CHLA-PLM07628564/2020\|EPI_ISL_75355 | ···················· | ···················· | ····················· | ···················· | ···················· | ···················· |
| AA | 4548 hCoV-19/USA/CA-CHLA-PLM29469835/2020\|EPI_ISL_75355 | ···················· | ···················· | ····················· | ···················· | ···················· | ···················· |
| AA | 4549 hCoV-19/USA/CA-CHLA-PLM21707363/2020\|EPI_ISL_75355 | ···················· | ·A·················· | ····················· | ···················· | ···················· | ···················· |
| AA | 4550 hCoV-19/USA/CA-CHLA-PLM19042682/2020\|EPI_ISL_75355 | ···················· | ·A·················· | ····················· | ···················· | ···················· | ···················· |
| AA | 4551 hCoV-19/USA/CA-CHLA-PLM28392841/2020\|EPI_ISL_75355 | ···················· | ···················· | ····················· | ···················· | ···················· | ···················· |
| AA | 4552 hCoV-19/USA/CA-CHLA-PLM99693185/2020\|EPI_ISL_75355 | ···················· | ·A·················· | ····················· | ···················· | ···················· | ···················· |
| AA | 4553 hCoV-19/USA/CA-CHLA-PLM15576410/2020\|EPI_ISL_75356 | ···················· | ···················· | ····················· | ···················· | ···················· | ···················· |
| AA | 4554 hCoV-19/USA/CA-CHLA-PLM87172348/2020\|EPI_ISL_75356 | ···················· | ···················· | ····················· | ···················· | ···················· | ···················· |
| AA | 4555 hCoV-19/USA/CA-CHLA-PLM41746412/2020\|EPI_ISL_75356 | ···················· | ···················· | ····················· | ···················· | ···················· | ···················· |
| AA | 4556 hCoV-19/USA/CA-CHLA-PLM68293216/2020\|EPI_ISL_75356 | ···················· | ···················· | ····················· | ···················· | ···················· | ···················· |
| AA | 4557 hCoV-19/USA/CA-CHLA-PLM51191431/2020\|EPI_ISL_75356 | ···················· | ···················· | ····················· | ···················· | ···················· | ···················· |
| AA | 4558 hCoV-19/USA/CA-CHLA-PLM29980142/2020\|EPI_ISL_75356 | ···················· | ·A·················· | ····················· | ···················· | ···················· | ···················· |
| AA | 4559 hCoV-19/USA/CA-CHLA-PLM08071546/2020\|EPI_ISL_75356 | ···················· | ···················· | ····················· | ···················· | ···················· | ···················· |
| AA | 4560 hCoV-19/USA/CA-CHLA-PLM48022068/2020\|EPI_ISL_75356 | ···················· | ···················· | ····················· | ···················· | ···················· | ···················· |
| AA | 4561 hCoV-19/USA/CA-CHLA-PLM21494482/2020\|EPI_ISL_75356 | ···················· | ···················· | ····················· | ···················· | ···················· | ···················· |
| AA | 4562 hCoV-19/USA/CA-CHLA-PLM97812099/2020\|EPI_ISL_75357 | ···················· | ···················· | ····················· | ···················· | ···················· | ···················· |
| AA | 4563 hCoV-19/USA/CA-CHLA-PLM22279170/2020\|EPI_ISL_75357 | ···················· | ·A·················· | ····················· | ···················· | ···················· | ···················· |

```
AA    4564 hCoV-19/USA/CA-CHLA-PLM55573502/2020|EPI_ISL_75357    ····················  ·A··················  ····················  ····················  ····················  ····················
AA    4565 hCoV-19/USA/CA-CHLA-PLM48917501/2020|EPI_ISL_75357    ····················  ·A··················  ····················  ····················  ····················  ····················
AA    4566 hCoV-19/USA/CA-CHLA-PLM14086335/2020|EPI_ISL_75357    ····················  ····················  ····················  ····················  ····················  ····················
AA    4567 hCoV-19/USA/CA-CHLA-PLM39513740/2020|EPI_ISL_75357    ····················  ····················  ····················  ····················  ····················  ····················
AA    4568 hCoV-19/USA/CA-CHLA-PLM97612587/2020|EPI_ISL_75357    ····················  ····················  ····················  ····················  ····················  ····················
AA    4569 hCoV-19/USA/CA-CHLA-PLM48652977/2020|EPI_ISL_75357    ····················  ·A··················  ····················  ····················  ····················  ····················
AA    4570 hCoV-19/USA/CA-CHLA-PLM20413192/2020|EPI_ISL_75357    ····················  ·A··················  ····················  ····················  ····················  ····················
AA    4571 hCoV-19/USA/NY-Wadsworth-251067-01/2020|EPI_ISL_75    ····················  ····················  ····················  ····················  ····················  ····················
AA    4572 hCoV-19/USA/NY-Wadsworth-251066-01/2020|EPI_ISL_75    ····················  ····················  ····················  ····················  ····················  ····················
AA    4573 hCoV-19/USA/NY-Wadsworth-250939-01/2020|EPI_ISL_75    ····················  ····················  ····················  ····················  ····················  ····················
AA    4574 hCoV-19/USA/NY-Wadsworth-250926-01/2020|EPI_ISL_75    ····················  ····················  ····················  ····················  ····················  ····················
AA    4575 hCoV-19/USA/CA-IGI-0325/2020|EPI_ISL_754866|2020-0    ····················  ····················  ····················  ····················  ····················  ····················
AA    4576 hCoV-19/USA/CA-IGI-0328/2020|EPI_ISL_754867|2020-0    ····················  ····················  ····················  ····················  ····················  ····················
AA    4577 hCoV-19/USA/CA-IGI-0329/2020|EPI_ISL_754868|2020-0    ····················  ····················  ····················  ····················  ····················  ····················
AA    4578 hCoV-19/USA/CA-IGI-0332/2020|EPI_ISL_754869|2020-0    ····················  ····················  ····················  ····················  ····················  ····················
AA    4579 hCoV-19/USA/CA-IGI-0334/2020|EPI_ISL_754870|2020-0    ····················  ·A··················  ····················  ····················  ····················  ····················
AA    4580 hCoV-19/USA/CA-IGI-0337/2020|EPI_ISL_754871|2020-0    ····················  ····················  ····················  ····················  ····················  ····················
AA    4581 hCoV-19/USA/CA-IGI-0343/2020|EPI_ISL_754872|2020-0    ····················  ·A··················  ····················  ····················  ····················  ····················
AA    4582 hCoV-19/USA/CA-IGI-0349/2020|EPI_ISL_754873|2020-0    ····················  ····················  ····················  ····················  ····················  ····················
AA    4583 hCoV-19/USA/CA-IGI-0350/2020|EPI_ISL_754874|2020-0    ····················  ·A··················  ····················  ····················  ····················  ····················
AA    4584 hCoV-19/USA/CA-IGI-0355/2020|EPI_ISL_754875|2020-0    ····················  ·········A··········  ····················  ····················  ····················  ····················
AA    4585 hCoV-19/USA/CA-IGI-0366/2020|EPI_ISL_754876|2020-0    ····················  ····················  ····················  ····················  ····················  ····················
AA    4586 hCoV-19/USA/CA-IGI-0371/2020|EPI_ISL_754877|2020-0    ····················  ····················  ····················  ····················  ····················  ····················
AA    4587 hCoV-19/USA/CA-IGI-0385/2020|EPI_ISL_754878|2020-0    ····················  ····················  ····················  ····················  ····················  ····················
AA    4588 hCoV-19/USA/CA-IGI-0386/2020|EPI_ISL_754879|2020-0    ····················  ·A··················  ····················  ····················  ····················  ····················
AA    4589 hCoV-19/USA/CA-IGI-0388/2020|EPI_ISL_754880|2020-0    ····················  ·A··················  ····················  ····················  ····················  ····················
AA    4590 hCoV-19/USA/CA-IGI-0391/2020|EPI_ISL_754881|2020-0    ····················  ····················  ····················  ····················  ····················  ····················
AA    4591 hCoV-19/USA/CA-IGI-0392/2020|EPI_ISL_754882|2020-0    ····················  ····················  ····················  ····················  ····················  ····················
AA    4592 hCoV-19/USA/CA-IGI-0394/2020|EPI_ISL_754883|2020-0    ····················  ····················  ····················  ····················  ····················  ····················
AA    4593 hCoV-19/USA/CA-IGI-0397/2020|EPI_ISL_754884|2020-0    ····················  ····················  ····················  ····················  ····················  ····················
AA    4594 hCoV-19/USA/CA-IGI-0400/2020|EPI_ISL_754885|2020-0    ····················  ····················  ····················  ····················  ····················  ····················
AA    4595 hCoV-19/USA/CA-IGI-0402/2020|EPI_ISL_754886|2020-0    ····················  ····················  ····················  ····················  ····················  ····················
AA    4596 hCoV-19/USA/CA-IGI-0404/2020|EPI_ISL_754887|2020-0    ····················  ·············A······  ····················  ····················  ····················  ····················
AA    4597 hCoV-19/USA/CA-IGI-0405/2020|EPI_ISL_754888|2020-0    ····················  ·A··················  ····················  ····················  ····················  ····················
AA    4598 hCoV-19/USA/CA-IGI-0407/2020|EPI_ISL_754889|2020-0    ····················  ····················  ····················  ····················  ····················  ····················
AA    4599 hCoV-19/USA/CA-IGI-0409/2020|EPI_ISL_754890|2020-0    ····················  ····················  ····················  ····················  ····················  ····················
AA    4600 hCoV-19/USA/CA-IGI-0410/2020|EPI_ISL_754891|2020-0    ····················  ····················  ····················  ····················  ····················  ····················
AA    4601 hCoV-19/USA/CA-IGI-0411/2020|EPI_ISL_754892|2020-0    ····················  ····················  ····················  ····················  ····················  ····················
AA    4602 hCoV-19/USA/CA-IGI-0413/2020|EPI_ISL_754893|2020-0    ····················  ····················  ····················  ····················  ····················  ····················
AA    4603 hCoV-19/USA/CA-IGI-0415/2020|EPI_ISL_754894|2020-0    ····················  ····················  ····················  ····················  ····················  ····················
AA    4604 hCoV-19/USA/CA-IGI-0416/2020|EPI_ISL_754895|2020-0    ····················  ·A··················  ····················  ····················  ····················  ····················
AA    4605 hCoV-19/USA/CA-IGI-0418/2020|EPI_ISL_754896|2020-0    ····················  ·A··················  ····················  ····················  ····················  ····················
AA    4606 hCoV-19/USA/CA-IGI-0422/2020|EPI_ISL_754897|2020-0    ····················  ····A···············  ····················  ····················  ····················  ····················
AA    4607 hCoV-19/USA/CA-IGI-0426/2020|EPI_ISL_754898|2020-0    ····················  ····················  ····················  ····················  ····················  ····················
AA    4608 hCoV-19/USA/CA-IGI-0431/2020|EPI_ISL_754899|2020-0    ····················  ····················  ····················  ····················  ····················  ····················
AA    4609 hCoV-19/Denmark/DCGC-20036/2020|EPI_ISL_747700|202    ····················  ····················  ····················  ····················  ····················  ····················
AA    4610 hCoV-19/Denmark/DCGC-20039/2020|EPI_ISL_747701|202    ····················  ····················  ····················  ····················  ····················  ····················
AA    4611 hCoV-19/Denmark/DCGC-20046/2020|EPI_ISL_747702|202    ····················  ····················  ····················  ····················  ····················  ····················
AA    4612 hCoV-19/Denmark/DCGC-20072/2020|EPI_ISL_747703|202    ····················  ····················  ····················  ····················  ····················  ····················
AA    4613 hCoV-19/Denmark/DCGC-20073/2020|EPI_ISL_747704|202    ····················  ····················  ····················  ····················  ····················  ····················
AA    4614 hCoV-19/Denmark/DCGC-20082/2020|EPI_ISL_747705|202    ····················  ····················  ····················  ····················  ····················  ····················
AA    4615 hCoV-19/Denmark/DCGC-20091/2020|EPI_ISL_747706|202    ····················  ····················  ····················  ····················  ····················  ····················
AA    4616 hCoV-19/Denmark/DCGC-20140/2020|EPI_ISL_747707|202    ····················  ····················  ····················  ····················  ····················  ····················
AA    4617 hCoV-19/Denmark/DCGC-20145/2020|EPI_ISL_747708|202    ····················  ····················  ····················  ····················  ····················  ····················
AA    4618 hCoV-19/Denmark/DCGC-20174/2020|EPI_ISL_747709|202    ····················  ····················  ····················  ····················  ····················  ····················
AA    4619 hCoV-19/Denmark/DCGC-20189/2020|EPI_ISL_747710|202    ····················  ····················  ····················  ····················  ····················  ····················
AA    4620 hCoV-19/Denmark/DCGC-20198/2020|EPI_ISL_747711|202    ····················  ····················  ····················  ····················  ····················  ····················
AA    4621 hCoV-19/Denmark/DCGC-20209/2020|EPI_ISL_747712|202    ····················  ····················  ····················  ····················  ····················  ····················
AA    4622 hCoV-19/Denmark/DCGC-20211/2020|EPI_ISL_747713|202    ····················  ····················  ····················  ····················  ····················  ····················
AA    4623 hCoV-19/Denmark/DCGC-20213/2020|EPI_ISL_747714|202    ····················  ····················  ····················  ····················  ····T···············  ····················
AA    4624 hCoV-19/Denmark/DCGC-20222/2020|EPI_ISL_747715|202    ····················  ····················  ····················  ····················  ····················  ····················
AA    4625 hCoV-19/Denmark/DCGC-20229/2020|EPI_ISL_747716|202    ····················  ····················  ····················  ····················  ····················  ····················
AA    4626 hCoV-19/Denmark/DCGC-20230/2020|EPI_ISL_747717|202    ····················  ····················  ····················  ····················  ····················  ····················
AA    4627 hCoV-19/Denmark/DCGC-20269/2020|EPI_ISL_747718|202    ····················  ····················  ····················  ····················  ····················  ····················
AA    4628 hCoV-19/Denmark/DCGC-20279/2020|EPI_ISL_747719|202    ····················  ····················  ····················  ····················  ····················  ····················
AA    4629 hCoV-19/Denmark/DCGC-20321/2020|EPI_ISL_747720|202    ····················  ····················  ····················  ····················  ····················  ····················
AA    4630 hCoV-19/Denmark/DCGC-19799/2020|EPI_ISL_747728|202    ····················  ····················  ····················  ····················  ····················  ····················
AA    4631 hCoV-19/Denmark/DCGC-19901/2020|EPI_ISL_747729|202    ····················  ·A··················  ····················  ····················  ····················  ···A················
AA    4632 hCoV-19/Denmark/DCGC-20259/2020|EPI_ISL_747730|202    ····················  ····················  ····················  ····················  ····················  ····················
AA    4633 hCoV-19/Denmark/DCGC-19749/2020|EPI_ISL_747756|202    ····················  ····················  ····················  ····················  ····················  ····················
AA    4634 hCoV-19/Denmark/DCGC-19750/2020|EPI_ISL_747757|202    ····················  ····················  ····················  ····················  ····················  ····················
AA    4635 hCoV-19/Denmark/DCGC-19751/2020|EPI_ISL_747758|202    ····················  ····················  ····················  ····················  ····················  ····················
AA    4636 hCoV-19/Denmark/DCGC-19753/2020|EPI_ISL_747759|202    ····················  ····················  ····················  ····················  ····················  ····················
AA    4637 hCoV-19/Denmark/DCGC-19754/2020|EPI_ISL_747760|202    ····················  ····················  ····················  ····················  ····················  ····················
AA    4638 hCoV-19/Denmark/DCGC-19755/2020|EPI_ISL_747761|202    ····················  ····················  ····················  ····················  ····················  ····················
AA    4639 hCoV-19/Denmark/DCGC-19756/2020|EPI_ISL_747762|202    ····················  ····················  ····················  ····················  ····················  ····················
AA    4640 hCoV-19/Denmark/DCGC-19757/2020|EPI_ISL_747763|202    ····················  ····················  ····················  ····················  ····················  ····················
AA    4641 hCoV-19/Denmark/DCGC-19758/2020|EPI_ISL_747764|202    ····················  ····················  ····················  ····················  ····················  ····················
AA    4642 hCoV-19/Denmark/DCGC-19759/2020|EPI_ISL_747765|202    ····················  ····················  ····················  ····················  ····················  ····················
AA    4643 hCoV-19/Denmark/DCGC-19760/2020|EPI_ISL_747766|202    ····················  ·············A······  ····················  ····················  ····················  ····················
```

| | | | | | | | |
|---|---|---|---|---|---|---|---|
| AA | 4644 hCoV-19/Denmark/DCGC-19761/2020\|EPI_ISL_747767\|202 | ···················· | ···················· | ···················· | ···················· | ···················· | ···················· |
| AA | 4645 hCoV-19/Denmark/DCGC-19762/2020\|EPI_ISL_747768\|202 | ···················· | ···················· | ···················· | ···················· | ···················· | ···················· |
| AA | 4646 hCoV-19/Denmark/DCGC-19763/2020\|EPI_ISL_747769\|202 | ···················· | ···················· | ···················· | ···················· | ···················· | ···················· |
| AA | 4647 hCoV-19/Denmark/DCGC-19764/2020\|EPI_ISL_747770\|202 | ···················· | ···················· | ···················· | ···················· | ···················· | ···················· |
| AA | 4648 hCoV-19/Denmark/DCGC-19766/2020\|EPI_ISL_747771\|202 | ···················· | ···················· | ···················· | ···················· | ···················· | ···················· |
| AA | 4649 hCoV-19/Denmark/DCGC-19767/2020\|EPI_ISL_747772\|202 | ···················· | ···················· | ···················· | ···················· | ···················· | ···················· |
| AA | 4650 hCoV-19/Denmark/DCGC-19768/2020\|EPI_ISL_747773\|202 | ···················· | ···················· | ···················· | ···················· | ···················· | ···················· |
| AA | 4651 hCoV-19/Denmark/DCGC-19769/2020\|EPI_ISL_747774\|202 | ···················· | ···················· | ···················· | ···················· | ···················· | ···················· |
| AA | 4652 hCoV-19/Denmark/DCGC-19770/2020\|EPI_ISL_747775\|202 | ···················· | ···················· | ···················· | ···················· | ···················· | ···················· |
| AA | 4653 hCoV-19/Denmark/DCGC-19771/2020\|EPI_ISL_747776\|202 | ···················· | ···················· | ···················· | ···················· | ···················· | ···················· |
| AA | 4654 hCoV-19/Denmark/DCGC-19772/2020\|EPI_ISL_747777\|202 | ···················· | ···················· | ···················· | ···················· | ···················· | ···················· |
| AA | 4655 hCoV-19/Denmark/DCGC-19773/2020\|EPI_ISL_747778\|202 | ···················· | ···················· | ···················· | ···················· | ···················· | ···················· |
| AA | 4656 hCoV-19/Denmark/DCGC-19774/2020\|EPI_ISL_747779\|202 | ···················· | ···················· | ···················· | ···················· | ···················· | ···················· |
| AA | 4657 hCoV-19/USA/CA-CHLA-PLM55178805/2020\|EPI_ISL_75350 | ···················· | ···················· | ···················· | ···················· | ···················· | ···················· |
| AA | 4658 hCoV-19/USA/CA-CHLA-PLM33320225/2020\|EPI_ISL_75350 | ···················· | ···················· | ···················· | ···················· | ···················· | ···················· |
| AA | 4659 hCoV-19/USA/CA-CHLA-PLM03408589/2020\|EPI_ISL_75350 | ···················· | ···················· | ···················· | ···················· | ···················· | ···················· |
| AA | 4660 hCoV-19/USA/CA-CHLA-PLM48846138/2020\|EPI_ISL_75350 | ···················· | ···················· | ···················· | ···················· | ···················· | ···················· |
| AA | 4661 hCoV-19/USA/CA-CHLA-PLM54539342/2020\|EPI_ISL_75350 | ···················· | ···················· | ···················· | ···················· | ···················· | ···················· |
| AA | 4662 hCoV-19/USA/CA-CHLA-PLM54662360/2020\|EPI_ISL_75350 | ···················· | ···················· | ···················· | ···················· | ···················· | ···················· |
| AA | 4663 hCoV-19/USA/CA-CHLA-PLM37711096/2020\|EPI_ISL_75350 | ···················· | ···················· | ···················· | ···················· | ···················· | ···················· |
| AA | 4664 hCoV-19/USA/CA-CHLA-PLM90708228/2020\|EPI_ISL_75350 | ···················· | ···················· | ···················· | ···················· | ···················· | ···················· |
| AA | 4665 hCoV-19/USA/CA-CHLA-PLM88767481/2020\|EPI_ISL_75350 | ···················· | ···················· | ···················· | ···················· | ···················· | ···················· |
| AA | 4666 hCoV-19/USA/CA-CHLA-PLM54461605/2020\|EPI_ISL_75350 | ···················· | ···················· | ···················· | ···················· | ···················· | ···················· |
| AA | 4667 hCoV-19/USA/CA-CHLA-PLM57308775/2020\|EPI_ISL_75351 | ···················· | ···················· | ···················· | ···················· | ···················· | ···················· |
| AA | 4668 hCoV-19/USA/CA-CHLA-PLM95086348/2020\|EPI_ISL_75351 | ···················· | ···················· | ···················· | ···················· | ···················· | ···················· |
| AA | 4669 hCoV-19/USA/CA-CHLA-PLM57076996/2020\|EPI_ISL_75351 | ···················· | ·A·················· | ···················· | ···················· | ···················· | ···················· |
| AA | 4670 hCoV-19/USA/CA-CHLA-PLM51977678/2020\|EPI_ISL_75351 | ···················· | ···················· | ···················· | ···················· | ···················· | ···················· |
| AA | 4671 hCoV-19/USA/CA-CHLA-PLM66830386/2020\|EPI_ISL_75351 | ···················· | ·A·················· | ···················· | ···················· | ···················· | ···················· |
| AA | 4672 hCoV-19/USA/CA-CHLA-PLM58014251/2020\|EPI_ISL_75351 | ···················· | ···················· | ···················· | ···················· | ···················· | ···················· |
| AA | 4673 hCoV-19/USA/CA-CHLA-PLM41636829/2020\|EPI_ISL_75351 | ···················· | ·A·················· | ···················· | ···················· | ···················· | ···················· |
| AA | 4674 hCoV-19/USA/CA-CHLA-PLM55758128/2020\|EPI_ISL_75351 | ···················· | ···················· | ···················· | ···················· | ···················· | ···················· |
| AA | 4675 hCoV-19/USA/CA-CHLA-PLM77922729/2020\|EPI_ISL_75351 | ···················· | ·A·················· | ···················· | ···················· | ···················· | ···················· |
| AA | 4676 hCoV-19/USA/CA-CHLA-PLM19259147/2020\|EPI_ISL_75351 | ···················· | ···················· | ···················· | ···················· | ···················· | ···················· |
| AA | 4677 hCoV-19/USA/CA-CHLA-PLM82326358/2020\|EPI_ISL_75352 | ···················· | ···················· | ···················· | ···················· | ···················· | ···················· |
| AA | 4678 hCoV-19/USA/CA-CHLA-PLM38778784/2020\|EPI_ISL_75352 | ···················· | ···················· | ···················· | ···················· | ···················· | ···················· |
| AA | 4679 hCoV-19/USA/CA-CHLA-PLM35178516/2020\|EPI_ISL_75352 | ···················· | ·A·················· | ···················· | ···················· | ···················· | ···················· |
| AA | 4680 hCoV-19/USA/CA-CHLA-PLM59547698/2020\|EPI_ISL_75352 | ···················· | ···················· | ···················· | ···················· | ···················· | ···················· |
| AA | 4681 hCoV-19/USA/CA-CHLA-PLM21871523/2020\|EPI_ISL_75352 | ···················· | ···················· | ···················· | ···················· | ···················· | ···················· |
| AA | 4682 hCoV-19/USA/CA-CHLA-PLM86229141/2020\|EPI_ISL_75352 | ···················· | ···················· | ···················· | ···················· | ···················· | ···················· |
| AA | 4683 hCoV-19/USA/CA-CHLA-PLM56822120/2020\|EPI_ISL_75352 | ···················· | ···················· | ···················· | ···················· | ···················· | ···················· |
| AA | 4684 hCoV-19/USA/CA-CHLA-PLM28080902/2020\|EPI_ISL_75352 | ···················· | ·A·················· | ···················· | ···················· | ···················· | ···················· |
| AA | 4685 hCoV-19/USA/CA-CHLA-PLM75223157/2020\|EPI_ISL_75352 | ···················· | ···················· | ···················· | ···················· | ···················· | ···················· |
| AA | 4686 hCoV-19/USA/CA-CHLA-PLM50317441/2020\|EPI_ISL_75352 | ···················· | ·A·················· | ···················· | ···················· | ···················· | ···················· |
| AA | 4687 hCoV-19/USA/CA-CHLA-PLM59996756/2020\|EPI_ISL_75353 | ···················· | ·A·················· | ···················· | ···················· | ···················· | ···················· |
| AA | 4688 hCoV-19/USA/CA-CHLA-PLM07768214/2020\|EPI_ISL_75353 | ···················· | ···················· | ···················· | ···················· | ···················· | ···················· |
| AA | 4689 hCoV-19/USA/CA-CHLA-PLM25788503/2020\|EPI_ISL_75353 | ···················· | ···················· | ···················· | ···················· | ···················· | ···················· |
| AA | 4690 hCoV-19/USA/CA-CHLA-PLM61546012/2020\|EPI_ISL_75353 | ···················· | ···················· | ···················· | ···················· | ···················· | ···················· |
| AA | 4691 hCoV-19/USA/CA-CHLA-PLM50354444/2020\|EPI_ISL_75357 | ···················· | ···················· | ···················· | ···················· | ···················· | ···················· |
| AA | 4692 hCoV-19/USA/CA-CHLA-PLM97372016/2020\|EPI_ISL_75358 | ···················· | ···················· | ···················· | ···················· | ···················· | ···················· |
| AA | 4693 hCoV-19/USA/CA-CHLA-PLM70139370/2020\|EPI_ISL_75358 | ···················· | ···················· | ···················· | ···················· | ···················· | ···················· |
| AA | 4694 hCoV-19/USA/CA-CHLA-PLM52888374/2020\|EPI_ISL_75358 | ···················· | ···················· | ···················· | ···················· | ···················· | ···················· |
| AA | 4695 hCoV-19/USA/CA-CHLA-PLM16111404/2020\|EPI_ISL_75358 | ···················· | ·A·················· | ···················· | ···················· | ···················· | ···················· |
| AA | 4696 hCoV-19/USA/CA-CHLA-PLM07126518/2020\|EPI_ISL_75358 | ···················· | ·A·················· | ···················· | ···················· | ···················· | ···················· |
| AA | 4697 hCoV-19/USA/CA-CHLA-PLM65298749/2020\|EPI_ISL_75358 | ···················· | ···················· | ···················· | ···················· | ···················· | ···················· |
| AA | 4698 hCoV-19/USA/CA-CHLA-PLM40259119/2020\|EPI_ISL_75358 | ···················· | ···················· | ···················· | ···················· | ···················· | ···················· |
| AA | 4699 hCoV-19/USA/CA-CHLA-PLM53624288/2020\|EPI_ISL_75358 | ···················· | ···················· | ···················· | ···················· | ···················· | ···················· |
| AA | 4700 hCoV-19/USA/CA-CHLA-PLM20114271/2020\|EPI_ISL_75358 | ···················· | ···················· | ···················· | ···················· | ···················· | ···················· |
| AA | 4701 hCoV-19/USA/CA-CHLA-PLM73778085/2020\|EPI_ISL_75358 | ···················· | ···················· | ···················· | ···················· | ···················· | ···················· |
| AA | 4702 hCoV-19/USA/CA-CHLA-PLM04375107/2020\|EPI_ISL_75359 | ···················· | ···················· | ···················· | ···················· | ···················· | ···················· |
| AA | 4703 hCoV-19/USA/CA-CHLA-PLM46344829/2020\|EPI_ISL_75359 | ···················· | ···················· | ···················· | ···················· | ···················· | ···················· |
| AA | 4704 hCoV-19/USA/CA-CHLA-PLM11371375/2020\|EPI_ISL_75359 | ···················· | ·A·················· | ···················· | ···················· | ···················· | ···················· |
| AA | 4705 hCoV-19/USA/CA-CHLA-PLM94973155/2020\|EPI_ISL_75359 | ···················· | ·A·················· | ···················· | ···················· | ···················· | ···················· |
| AA | 4706 hCoV-19/USA/CA-CHLA-PLM32250719/2020\|EPI_ISL_75359 | ···················· | ·A·················· | ···················· | ···················· | ···················· | ···················· |
| AA | 4707 hCoV-19/USA/CA-CHLA-PLM45843117/2020\|EPI_ISL_75359 | ···················· | ···················· | ···················· | ···················· | ···················· | ···················· |
| AA | 4708 hCoV-19/USA/CA-CHLA-PLM50948447/2020\|EPI_ISL_75359 | ···················· | ···················· | ···················· | ···················· | ···················· | ···················· |
| AA | 4709 hCoV-19/USA/CA-CHLA-PLM26469289/2020\|EPI_ISL_75359 | ···················· | ···················· | ···················· | ···················· | ···················· | ···················· |
| AA | 4710 hCoV-19/USA/CA-CHLA-PLM60013986/2020\|EPI_ISL_75359 | ···················· | ···················· | ···················· | ···················· | ···················· | ···················· |
| AA | 4711 hCoV-19/USA/CA-CHLA-PLM37695791/2020\|EPI_ISL_75359 | ···················· | ·A·················· | ···················· | ···················· | ···················· | ···················· |
| AA | 4712 hCoV-19/USA/NY-Wadsworth-250999-01/2020\|EPI_ISL_75 | ···················· | ···················· | ···················· | ···················· | ···················· | ···················· |
| AA | 4713 hCoV-19/USA/NY-Wadsworth-251001-01/2020\|EPI_ISL_75 | ···················· | ···················· | ···················· | ···················· | ···················· | ···················· |
| AA | 4714 hCoV-19/USA/NY-Wadsworth-250982-01/2020\|EPI_ISL_75 | ···················· | ···················· | ···················· | ···················· | ···················· | ···················· |
| AA | 4715 hCoV-19/USA/NY-Wadsworth-250970-01/2020\|EPI_ISL_75 | ···················· | ···················· | ···················· | ···················· | ···················· | ···················· |
| AA | 4716 hCoV-19/USA/NY-Wadsworth-250969-01/2020\|EPI_ISL_75 | ···················· | ···················· | ···················· | ···················· | ···················· | ···················· |
| AA | 4717 hCoV-19/USA/NY-Wadsworth-251033-01/2020\|EPI_ISL_75 | ···················· | ···················· | ···················· | ···················· | ···················· | ···················· |
| AA | 4718 hCoV-19/USA/NY-Wadsworth-251029-01/2020\|EPI_ISL_75 | ···················· | ···················· | ···················· | ···················· | ···················· | ···················· |
| AA | 4719 hCoV-19/USA/NY-Wadsworth-250918-01/2020\|EPI_ISL_75 | ···················· | ···················· | ···················· | ···················· | ···················· | ···················· |
| AA | 4720 hCoV-19/USA/NY-Wadsworth-250959-01/2020\|EPI_ISL_75 | ···················· | ···················· | ···················· | ···················· | ···················· | ···················· |
| AA | 4721 hCoV-19/USA/NY-Wadsworth-250929-01/2020\|EPI_ISL_75 | ···················· | ···················· | ···················· | ···················· | ···················· | ···················· |
| AA | 4722 hCoV-19/USA/NY-Wadsworth-250928-01/2020\|EPI_ISL_75 | ···················· | ···················· | ···················· | ···················· | ···················· | ···················· |
| AA | 4723 hCoV-19/USA/NY-Wadsworth-251052-01/2020\|EPI_ISL_75 | ···················· | ···················· | ···················· | ···················· | ···················· | ···················· |

| | | | | | | | |
|---|---|---|---|---|---|---|---|
| AA | 4724 hCoV-19/USA/NY-Wadsworth-251011-01/2020\|EPI_ISL_75 | .................... | .................... | .................... | .................... | .................... | .................... |
| AA | 4725 hCoV-19/USA/NY-Wadsworth-250986-01/2020\|EPI_ISL_75 | .................... | .................... | .................... | .................... | .................... | .................... |
| AA | 4726 hCoV-19/USA/NY-Wadsworth-252077-01/2020\|EPI_ISL_75 | .................... | .................... | .................... | .................... | .................... | .................... |
| AA | 4727 hCoV-19/USA/NY-Wadsworth-252054-01/2020\|EPI_ISL_75 | .................... | .................... | .................... | .................... | .................... | .................... |
| AA | 4728 hCoV-19/France/BFC-IPP10929/2020\|EPI_ISL_754847\|20 | .................... | .................... | .................... | .................... | .................... | .................... |
| AA | 4729 hCoV-19/France/BFC-IPP10931/2020\|EPI_ISL_754848\|20 | .................... | .................... | .................... | .................... | .................... | .................... |
| AA | 4730 hCoV-19/France/GES-IPP10969/2020\|EPI_ISL_754849\|20 | .................... | .................... | .................... | .................... | .................... | .................... |
| AA | 4731 hCoV-19/France/GES-IPP10972/2020\|EPI_ISL_754850\|20 | .................... | .................... | .................... | .................... | .................... | .................... |
| AA | 4732 hCoV-19/France/GES-IPP10978/2020\|EPI_ISL_754851\|20 | .................... | .................... | .................... | .................... | .................... | .................... |
| AA | 4733 hCoV-19/France/GES-IPP10981/2020\|EPI_ISL_754852\|20 | .................... | .................... | .................... | .................... | .................... | .................... |
| AA | 4734 hCoV-19/France/BRE-IPP11307/2020\|EPI_ISL_754853\|20 | .................... | .................... | .................... | .................... | .................... | .................... |
| AA | 4735 hCoV-19/France/BRE-IPP11308/2020\|EPI_ISL_754854\|20 | .................... | .................... | .................... | .................... | .................... | .................... |
| AA | 4736 hCoV-19/France/BRE-IPP11309/2020\|EPI_ISL_754855\|20 | .................... | .................... | .................... | .................... | .................... | .................... |
| AA | 4737 hCoV-19/France/BRE-IPP11310/2020\|EPI_ISL_754856\|20 | .................... | .................... | .................... | .................... | .................... | .................... |
| AA | 4738 hCoV-19/USA/CA-IGI-0399/2020\|EPI_ISL_754864\|2020-0 | .................... | .................... | .................... | .................... | .................... | .................... |
| AA | 4739 hCoV-19/USA/CA-IGI-0322/2020\|EPI_ISL_754865\|2020-0 | .................... | .................... | .................... | .................... | .................... | .................... |
| AA | 4740 hCoV-19/Scotland/QEUH-C4BF00/2020\|EPI_ISL_757100\|2 | .................... | .................... | .................... | .................... | .................... | .................... |
| AA | 4741 hCoV-19/Scotland/QEUH-C4BC72/2020\|EPI_ISL_757101\|2 | .................... | .................... | .................... | .................... | .................... | .................... |
| AA | 4742 hCoV-19/Scotland/QEUH-C4BC27/2020\|EPI_ISL_757102\|2 | .................... | .................... | .................... | .................... | .................... | .................... |
| AA | 4743 hCoV-19/Scotland/QEUH-C4BC54/2020\|EPI_ISL_757104\|2 | .................... | .................... | .................... | .................... | .................... | .................... |
| AA | 4744 hCoV-19/Scotland/QEUH-C4BFD3/2020\|EPI_ISL_757105\|2 | .................... | .................... | .................... | .................... | .................... | .................... |
| AA | 4745 hCoV-19/Scotland/QEUH-C4C066/2020\|EPI_ISL_757107\|2 | .................... | .................... | .................... | .................... | .................... | .................... |
| AA | 4746 hCoV-19/Scotland/QEUH-C4C01B/2020\|EPI_ISL_757111\|2 | .................... | .................... | .................... | .................... | .................... | .................... |
| AA | 4747 hCoV-19/Scotland/QEUH-C4BE8B/2020\|EPI_ISL_757115\|2 | .................... | .................... | .................... | .................... | .................... | .................... |
| AA | 4748 hCoV-19/Scotland/QEUH-C4BF2E/2020\|EPI_ISL_757117\|2 | .................... | .................... | .................... | .................... | .................... | .................... |
| AA | 4749 hCoV-19/Scotland/QEUH-C4BC36/2020\|EPI_ISL_757118\|2 | .................... | .................... | .................... | .................... | .................... | .................... |
| AA | 4750 hCoV-19/Scotland/QEUH-C30FA4/2020\|EPI_ISL_757120\|2 | .................... | .................... | .................... | .................... | .................... | .................... |
| AA | 4751 hCoV-19/Scotland/QEUH-C319F0/2020\|EPI_ISL_757121\|2 | .................... | .................... | .................... | .................... | .................... | .................... |
| AA | 4752 hCoV-19/Scotland/QEUH-C30FD1/2020\|EPI_ISL_757122\|2 | .................... | .................... | .................... | .................... | .................... | .................... |
| AA | 4753 hCoV-19/Scotland/QEUH-C30D04/2020\|EPI_ISL_757123\|2 | .................... | .................... | .................... | .................... | .................... | .................... |
| AA | 4754 hCoV-19/Scotland/QEUH-C30C8F/2020\|EPI_ISL_757124\|2 | .................... | .................... | .................... | .................... | .................... | .................... |
| AA | 4755 hCoV-19/Scotland/QEUH-C30CE9/2020\|EPI_ISL_757125\|2 | .................... | .................... | .................... | .................... | .................... | .................... |
| AA | 4756 hCoV-19/Scotland/QEUH-C30F86/2020\|EPI_ISL_757126\|2 | .................... | .................... | .................... | .................... | .................... | .................... |
| AA | 4757 hCoV-19/Scotland/QEUH-C30D13/2020\|EPI_ISL_757127\|2 | .................... | .................... | .................... | .................... | .................... | .................... |
| AA | 4758 hCoV-19/Scotland/QEUH-C30CAD/2020\|EPI_ISL_757128\|2 | .................... | .................... | .................... | .................... | .................... | .................... |
| AA | 4759 hCoV-19/Scotland/QEUH-C30D31/2020\|EPI_ISL_757129\|2 | .................... | .................... | .................... | .................... | .................... | .................... |
| AA | 4760 hCoV-19/Scotland/QEUH-C30C9E/2020\|EPI_ISL_757130\|2 | .................... | .................... | .................... | .................... | .................... | .................... |
| AA | 4761 hCoV-19/Scotland/QEUH-C30CCB/2020\|EPI_ISL_757131\|2 | .................... | .................... | .................... | .................... | .................... | .................... |
| AA | 4762 hCoV-19/Scotland/QEUH-C30C61/2020\|EPI_ISL_757132\|2 | .................... | .................... | .................... | .................... | .................... | .................... |
| AA | 4763 hCoV-19/Scotland/QEUH-C30CF8/2020\|EPI_ISL_757133\|2 | .................... | .................... | .................... | .................... | .................... | .................... |
| AA | 4764 hCoV-19/Scotland/QEUH-C30C70/2020\|EPI_ISL_757134\|2 | .................... | .................... | .................... | .................... | .................... | .................... |
| AA | 4765 hCoV-19/Scotland/QEUH-C30CDA/2020\|EPI_ISL_757135\|2 | .................... | .................... | .................... | .................... | .................... | .................... |
| AA | 4766 hCoV-19/Scotland/QEUH-C3432C/2020\|EPI_ISL_757136\|2 | .................... | .................... | .................... | .................... | .................... | .................... |
| AA | 4767 hCoV-19/Scotland/QEUH-C33651/2020\|EPI_ISL_757137\|2 | .................... | .................... | .................... | .................... | .................... | .................... |
| AA | 4768 hCoV-19/Scotland/QEUH-C33C31/2020\|EPI_ISL_757138\|2 | .................... | .................... | .................... | .................... | .................... | .................... |
| AA | 4769 hCoV-19/Scotland/QEUH-C33BE9/2020\|EPI_ISL_757139\|2 | .................... | .................... | .................... | .................... | .................... | .................... |
| AA | 4770 hCoV-19/Denmark/DCGC-21845/2020\|EPI_ISL_758400\|202 | .................... | .................... | .................... | .................... | .................... | .................... |
| AA | 4771 hCoV-19/Denmark/DCGC-21846/2020\|EPI_ISL_758401\|202 | .................... | .................... | .................... | .................... | .................... | .................... |
| AA | 4772 hCoV-19/Denmark/DCGC-21848/2020\|EPI_ISL_758402\|202 | .................... | .................... | .................... | .................... | .................... | .................... |
| AA | 4773 hCoV-19/Denmark/DCGC-21849/2020\|EPI_ISL_758403\|202 | .................... | .................... | .................... | .................... | .................... | .................... |
| AA | 4774 hCoV-19/Denmark/DCGC-21853/2020\|EPI_ISL_758406\|202 | .................... | .................... | .................... | .................... | .................... | .................... |
| AA | 4775 hCoV-19/Denmark/DCGC-21854/2020\|EPI_ISL_758407\|202 | .................... | .................... | .................... | .................... | .................... | .................... |
| AA | 4776 hCoV-19/Denmark/DCGC-21884/2020\|EPI_ISL_758430\|202 | .................... | .................... | .................... | .................... | .................... | .................... |
| AA | 4777 hCoV-19/Denmark/DCGC-21886/2020\|EPI_ISL_758431\|202 | .................... | .................... | .................... | .................... | .................... | .................... |
| AA | 4778 hCoV-19/Denmark/DCGC-21887/2020\|EPI_ISL_758432\|202 | .................... | .................... | .................... | .................... | .................... | .................... |
| AA | 4779 hCoV-19/Denmark/DCGC-21888/2020\|EPI_ISL_758433\|202 | .................... | .................... | .................... | .................... | .................... | .................... |
| AA | 4780 hCoV-19/Denmark/DCGC-21889/2020\|EPI_ISL_758434\|202 | .................... | .................... | .................... | .................... | .................... | .................... |
| AA | 4781 hCoV-19/Denmark/DCGC-21890/2020\|EPI_ISL_758435\|202 | .................... | .................... | .................... | .................... | .................... | .................... |
| AA | 4782 hCoV-19/Denmark/DCGC-21891/2020\|EPI_ISL_758436\|202 | .................... | .................... | .................... | .................... | .................... | .................... |
| AA | 4783 hCoV-19/Denmark/DCGC-21894/2020\|EPI_ISL_758437\|202 | .................... | .................... | .................... | .................... | .................... | .................... |
| AA | 4784 hCoV-19/Denmark/DCGC-21895/2020\|EPI_ISL_758438\|202 | .................... | .................... | .................... | .................... | .................... | .................... |
| AA | 4785 hCoV-19/Denmark/DCGC-21896/2020\|EPI_ISL_758439\|202 | .................... | .................... | .................... | .................... | .................... | .................... |
| AA | 4786 hCoV-19/Denmark/DCGC-21898/2020\|EPI_ISL_758440\|202 | .................... | .................... | .................... | .................... | .................... | .................... |
| AA | 4787 hCoV-19/Denmark/DCGC-21899/2020\|EPI_ISL_758441\|202 | .................... | .................... | .................... | .................... | .................... | .................... |
| AA | 4788 hCoV-19/Denmark/DCGC-21900/2020\|EPI_ISL_758442\|202 | .................... | .................... | .................... | .................... | .................... | .................... |
| AA | 4789 hCoV-19/Denmark/DCGC-21901/2020\|EPI_ISL_758443\|202 | .................... | .................... | .................... | .................... | .................... | .................... |
| AA | 4790 hCoV-19/Denmark/DCGC-21902/2020\|EPI_ISL_758444\|202 | .................... | .................... | .................... | .................... | .................... | .................... |
| AA | 4791 hCoV-19/Denmark/DCGC-21903/2020\|EPI_ISL_758445\|202 | .................... | .................... | .................... | .................... | .................... | .................... |
| AA | 4792 hCoV-19/Denmark/DCGC-21904/2020\|EPI_ISL_758446\|202 | .................... | .................... | .................... | .................... | .................... | .................... |
| AA | 4793 hCoV-19/Denmark/DCGC-21906/2020\|EPI_ISL_758447\|202 | .................... | .................... | .................... | .................... | .................... | .................... |
| AA | 4794 hCoV-19/Denmark/DCGC-21908/2020\|EPI_ISL_758448\|202 | .................... | .................... | .................... | .................... | .................... | .................... |
| AA | 4795 hCoV-19/Denmark/DCGC-21909/2020\|EPI_ISL_758449\|202 | .................... | .................... | .................... | .................... | .................... | .................... |
| AA | 4796 hCoV-19/Denmark/DCGC-21910/2020\|EPI_ISL_758450\|202 | .................... | .................... | .................... | .................... | .................... | .................... |
| AA | 4797 hCoV-19/Denmark/DCGC-21911/2020\|EPI_ISL_758451\|202 | .................... | .................... | .................... | .................... | .................... | .................... |
| AA | 4798 hCoV-19/Denmark/DCGC-21913/2020\|EPI_ISL_758452\|202 | .................... | .................... | .................... | .................... | .................... | .................... |
| AA | 4799 hCoV-19/Denmark/DCGC-21914/2020\|EPI_ISL_758453\|202 | .................... | .................... | .................... | .................... | .................... | .................... |
| AA | 4800 hCoV-19/Denmark/DCGC-21915/2020\|EPI_ISL_758454\|202 | .................... | .................... | .................... | .................... | .................... | .................... |
| AA | 4801 hCoV-19/Denmark/DCGC-21917/2020\|EPI_ISL_758455\|202 | .................... | .................... | .................... | .................... | .................... | .................... |
| AA | 4802 hCoV-19/Denmark/DCGC-21919/2020\|EPI_ISL_758456\|202 | .................... | .................... | .................... | .................... | .................... | .................... |
| AA | 4803 hCoV-19/Denmark/DCGC-21920/2020\|EPI_ISL_758457\|202 | .................... | .................... | .................... | .................... | .................... | .................... |

| | | | | | | | |
|---|---|---|---|---|---|---|---|
| AA | 4804 hCoV-19/Denmark/DCGC-21921/2020\|EPI_ISL_758458\|202 | ···················· | ···················· | ···················· | ···················· | ···················· | ···················· |
| AA | 4805 hCoV-19/Denmark/DCGC-21923/2020\|EPI_ISL_758459\|202 | ···················· | ···················· | ···················· | ···················· | ···················· | ···················· |
| AA | 4806 hCoV-19/Denmark/DCGC-21928/2020\|EPI_ISL_758460\|202 | ···················· | ···················· | ···················· | ···················· | ···················· | ···················· |
| AA | 4807 hCoV-19/Denmark/DCGC-21929/2020\|EPI_ISL_758461\|202 | ···················· | ···················· | ···················· | ···················· | ···················· | ···················· |
| AA | 4808 hCoV-19/Denmark/DCGC-21931/2020\|EPI_ISL_758462\|202 | ···················· | ·A·················· | ···················· | ···················· | ···················· | ··A················· |
| AA | 4809 hCoV-19/Denmark/DCGC-21936/2020\|EPI_ISL_758463\|202 | ···················· | ···················· | ···················· | ···················· | ···················· | ···················· |
| AA | 4810 hCoV-19/Denmark/DCGC-21938/2020\|EPI_ISL_758464\|202 | ···················· | ···················· | ···················· | ···················· | ···················· | ···················· |
| AA | 4811 hCoV-19/Denmark/DCGC-21939/2020\|EPI_ISL_758465\|202 | ···················· | ···················· | ···················· | ···················· | ···················· | ···················· |
| AA | 4812 hCoV-19/Denmark/DCGC-21940/2020\|EPI_ISL_758466\|202 | ···················· | ···················· | ···················· | ···················· | ···················· | ···················· |
| AA | 4813 hCoV-19/Denmark/DCGC-21943/2020\|EPI_ISL_758467\|202 | ···················· | ···················· | ···················· | ···················· | ···················· | ···················· |
| AA | 4814 hCoV-19/Denmark/DCGC-21944/2020\|EPI_ISL_758468\|202 | ···················· | ···················· | ···················· | ···················· | ···················· | ···················· |
| AA | 4815 hCoV-19/Denmark/DCGC-21945/2020\|EPI_ISL_758469\|202 | ···················· | ···················· | ···················· | ···················· | ···················· | ···················· |
| AA | 4816 hCoV-19/Denmark/DCGC-21358/2020\|EPI_ISL_759700\|202 | ···················· | ···················· | ···················· | ···················· | ···················· | ···················· |
| AA | 4817 hCoV-19/Indonesia/JT-UNS-18_39701/2020\|EPI_ISL_759 | ···················· | ···················· | ···················· | ···················· | ···················· | ···················· |
| AA | 4818 hCoV-19/Portugal/PT2128/2020\|EPI_ISL_759719\|2020-1 | ···················· | ···················· | ···················· | ···················· | ···················· | ···················· |
| AA | 4820 hCoV-19/USA/WI-UW-2450/2020\|EPI_ISL_759725\|2020-12 | ···················· | ···················· | ···················· | ···················· | ···················· | ···················· |
| AA | 4822 hCoV-19/USA/WI-UW-2453/2020\|EPI_ISL_759728\|2020-12 | ···················· | ···················· | ···················· | ···················· | ···················· | ···················· |
| AA | 4823 hCoV-19/USA/WI-UW-2454/2020\|EPI_ISL_759729\|2020-12 | ···················· | ···················· | ···················· | ···················· | ···················· | ···················· |
| AA | 4824 hCoV-19/USA/WI-UW-2456/2020\|EPI_ISL_759731\|2020-12 | ···················· | ···················· | ···················· | ···················· | ···················· | ···················· |
| AA | 4825 hCoV-19/USA/IA-UW-2458/2020\|EPI_ISL_759733\|2020-12 | ···················· | ···················· | ···················· | ···················· | ···················· | ···················· |
| AA | 4826 hCoV-19/USA/WI-UW-2459/2020\|EPI_ISL_759734\|2020-12 | ···················· | ···················· | ···················· | ···················· | ···················· | ···················· |
| AA | 4827 hCoV-19/USA/WI-UW-2462/2020\|EPI_ISL_759737\|2020-12 | ···················· | ···················· | ···················· | ···················· | ···················· | ···················· |
| AA | 4828 hCoV-19/Finland/23ES6AA5/2020\|EPI_ISL_759763\|2020- | ···················· | ···················· | ···················· | ···················· | ···················· | ···················· |
| AA | 4829 hCoV-19/Finland/23ES6AH5/2020\|EPI_ISL_759764\|2020- | ···················· | ···················· | ···················· | ···················· | ···················· | ···················· |
| AA | 4830 hCoV-19/Finland/23LS10H11/2020\|EPI_ISL_759765\|2020 | ···················· | ···················· | ···················· | ···················· | ···················· | ···················· |
| AA | 4831 hCoV-19/Finland/23LS11B7/2020\|EPI_ISL_759766\|2020- | ···················· | ·A·················· | ···················· | ···················· | ···················· | ··A················· |
| AA | 4832 hCoV-19/Finland/23LS17B11/2020\|EPI_ISL_759767\|2020 | ···················· | ·A·················· | ···················· | ···················· | ···················· | ··A················· |
| AA | 4833 hCoV-19/Finland/23LS17B5/2020\|EPI_ISL_759768\|2020- | ···················· | ·A·················· | ···················· | ···················· | ···················· | ··A················· |
| AA | 4834 hCoV-19/Finland/23LS17E11/2020\|EPI_ISL_759769\|2020 | ···················· | ···················· | ···················· | ···················· | ···················· | ···················· |
| AA | 4835 hCoV-19/Finland/23LS17H7/2020\|EPI_ISL_759770\|2020- | ···················· | ···················· | ···················· | ···················· | ···················· | ···················· |
| AA | 4836 hCoV-19/Finland/23LS1E6/2020\|EPI_ISL_759771\|2020-1 | ···················· | ···················· | ···················· | ···················· | ···················· | ···················· |
| AA | 4837 hCoV-19/Finland/23LS1F10/2020\|EPI_ISL_759772\|2020- | ···················· | ···················· | ···················· | ···················· | ···················· | ···················· |
| AA | 4838 hCoV-19/Finland/23LS1G7/2020\|EPI_ISL_759773\|2020-1 | ···················· | ···················· | ···················· | ···················· | ···················· | ···················· |
| AA | 4839 hCoV-19/Finland/23LS1H5/2020\|EPI_ISL_759774\|2020-1 | ···················· | ···················· | ···················· | ···················· | ···················· | ···················· |
| AA | 4840 hCoV-19/Finland/23LS3E11/2020\|EPI_ISL_759775\|2020- | ···················· | ···················· | ···················· | ···················· | ···················· | ···················· |
| AA | 4841 hCoV-19/Finland/23LS5C4/2020\|EPI_ISL_759776\|2020-1 | ···················· | ···················· | ···················· | ···················· | ···················· | ···················· |
| AA | 4842 hCoV-19/Finland/23LS5D9/2020\|EPI_ISL_759777\|2020-1 | ···················· | ···················· | ···················· | ···················· | ···················· | ···················· |
| AA | 4843 hCoV-19/Finland/23LS5G11/2020\|EPI_ISL_759778\|2020- | ···················· | ·A·················· | ···················· | ···················· | ···················· | ··A················· |
| AA | 4844 hCoV-19/Finland/23LS5H3/2020\|EPI_ISL_759779\|2020-1 | ···················· | ···················· | ···················· | ···················· | ···················· | ···················· |
| AA | 4845 hCoV-19/Finland/23LS6A12/2020\|EPI_ISL_759780\|2020- | ···················· | ·A·················· | ···················· | ···················· | ···················· | ··A················· |
| AA | 4846 hCoV-19/Finland/23LS6C11/2020\|EPI_ISL_759781\|2020- | ···················· | ·A·················· | ···················· | ···················· | ···················· | ··A················· |
| AA | 4847 hCoV-19/Finland/23LS6C12/2020\|EPI_ISL_759782\|2020- | ···················· | ·A·················· | ···················· | ···················· | ···················· | ··A················· |
| AA | 4848 hCoV-19/Finland/23LS6D12/2020\|EPI_ISL_759783\|2020- | ···················· | ·A·················· | ···················· | ···················· | ···················· | ··A················· |
| AA | 4849 hCoV-19/Finland/23LS6E9/2020\|EPI_ISL_759784\|2020-1 | ···················· | ·A·················· | ···················· | ···················· | ···················· | ··A················· |
| AA | 4850 hCoV-19/Finland/23LS6F7/2020\|EPI_ISL_759785\|2020-1 | ···················· | ·A·················· | ···················· | ···················· | ···················· | ··A················· |
| AA | 4851 hCoV-19/Finland/23LS6F8/2020\|EPI_ISL_759786\|2020-1 | ···················· | ·A·················· | ···················· | ···················· | ···················· | ··A················· |
| AA | 4852 hCoV-19/Finland/23LS6H10/2020\|EPI_ISL_759787\|2020- | ···················· | ·A·················· | ···················· | ···················· | ···················· | ··A················· |
| AA | 4853 hCoV-19/Finland/23LS7DD11/2020\|EPI_ISL_759788\|2020 | ···················· | ·A·················· | ···················· | ···················· | ···················· | ··A················· |
| AA | 4854 hCoV-19/Finland/23LS7DH3/2020\|EPI_ISL_759789\|2020- | ···················· | ·A·················· | ···················· | ···················· | ···················· | ··A················· |
| AA | 4855 hCoV-19/Finland/23LS7G2/2020\|EPI_ISL_759790\|2020-1 | ···················· | ·A·················· | ···················· | ···················· | ···················· | ··A················· |
| AA | 4856 hCoV-19/Finland/23LS8C1/2020\|EPI_ISL_759791\|2020-1 | ···················· | ·A·················· | ···················· | ···················· | ···················· | ··A················· |
| AA | 4857 hCoV-19/Finland/23LS8F3/2020\|EPI_ISL_759792\|2020-1 | ···················· | ·A·················· | ···················· | ···················· | ···················· | ··A················· |
| AA | 4858 hCoV-19/Finland/23LS8G1/2020\|EPI_ISL_759793\|2020-1 | ···················· | ·A·················· | ···················· | ···················· | ···················· | ··A················· |
| AA | 4859 hCoV-19/Finland/23LS9A7/2020\|EPI_ISL_759794\|2020-1 | ···················· | ···················· | ···················· | ···················· | ···················· | ···················· |
| AA | 4860 hCoV-19/Finland/23LS9H3/2020\|EPI_ISL_759795\|2020-1 | ···················· | ···················· | ···················· | ···················· | ···················· | ···················· |
| AA | 4861 hCoV-19/Finland/23SS10F11/2020\|EPI_ISL_759796\|2020 | ···················· | ·A·················· | ···················· | ···················· | ···················· | ··A················· |
| AA | 4862 hCoV-19/Finland/23SS11E11/2020\|EPI_ISL_759797\|2020 | ···················· | ·A·················· | ···················· | ···················· | ···················· | ··A················· |
| AA | 4863 hCoV-19/Finland/23SS12G8/2020\|EPI_ISL_759798\|2020- | ···················· | ·A·················· | ···················· | ···················· | ···················· | ··A················· |
| AA | 4864 hCoV-19/Finland/23SS13A5/2020\|EPI_ISL_759799\|2020- | ···················· | ·A·················· | ···················· | ···················· | ···················· | ··A················· |
| AA | 4865 hCoV-19/Denmark/DCGC-19775/2020\|EPI_ISL_747780\|202 | ···················· | ···················· | ···················· | ···················· | ···················· | ···················· |
| AA | 4866 hCoV-19/Denmark/DCGC-19777/2020\|EPI_ISL_747781\|202 | ···················· | ···················· | ···················· | ···················· | ···················· | ···················· |
| AA | 4867 hCoV-19/Denmark/DCGC-19778/2020\|EPI_ISL_747782\|202 | ···················· | ···················· | ···················· | ···················· | ···················· | ···················· |
| AA | 4868 hCoV-19/Denmark/DCGC-19779/2020\|EPI_ISL_747783\|202 | ···················· | ···················· | ···················· | ···················· | ···················· | ···················· |
| AA | 4869 hCoV-19/Denmark/DCGC-19780/2020\|EPI_ISL_747784\|202 | ···················· | ···················· | ···················· | ···················· | ···················· | ···················· |
| AA | 4870 hCoV-19/Denmark/DCGC-19782/2020\|EPI_ISL_747785\|202 | ···················· | ···················· | ···················· | ···················· | ···················· | ···················· |
| AA | 4871 hCoV-19/Denmark/DCGC-19784/2020\|EPI_ISL_747786\|202 | ···················· | ···················· | ···················· | ···················· | ···················· | ···················· |
| AA | 4872 hCoV-19/Denmark/DCGC-19786/2020\|EPI_ISL_747787\|202 | ···················· | ···················· | ···················· | ···················· | ···················· | ···················· |
| AA | 4873 hCoV-19/Denmark/DCGC-19788/2020\|EPI_ISL_747788\|202 | ···················· | ···················· | ···················· | ···················· | ···················· | ···················· |
| AA | 4874 hCoV-19/Denmark/DCGC-19789/2020\|EPI_ISL_747789\|202 | ···················· | ···················· | ···················· | ···················· | ···················· | ···················· |
| AA | 4875 hCoV-19/Denmark/DCGC-19790/2020\|EPI_ISL_747790\|202 | ···················· | ···················· | ···················· | ···················· | ···················· | ···················· |
| AA | 4876 hCoV-19/Denmark/DCGC-19791/2020\|EPI_ISL_747791\|202 | ···················· | ···················· | ···················· | ···················· | ···················· | ···················· |
| AA | 4877 hCoV-19/Denmark/DCGC-19792/2020\|EPI_ISL_747792\|202 | ···················· | ···················· | ···················· | ···················· | ···················· | ···················· |
| AA | 4878 hCoV-19/Denmark/DCGC-19796/2020\|EPI_ISL_747793\|202 | ···················· | ···················· | ···················· | ···················· | ···················· | ···················· |
| AA | 4879 hCoV-19/Denmark/DCGC-19797/2020\|EPI_ISL_747794\|202 | ···················· | ···················· | ···················· | ···················· | ···················· | ···················· |
| AA | 4880 hCoV-19/Denmark/DCGC-19798/2020\|EPI_ISL_747795\|202 | ···················· | ···················· | ···················· | ···················· | ···················· | ···················· |
| AA | 4881 hCoV-19/Denmark/DCGC-19800/2020\|EPI_ISL_747796\|202 | ···················· | ···················· | ···················· | ···················· | ···················· | ···················· |
| AA | 4882 hCoV-19/Denmark/DCGC-19801/2020\|EPI_ISL_747797\|202 | ···················· | ···················· | ···················· | ···················· | ···················· | ···················· |
| AA | 4883 hCoV-19/Denmark/DCGC-19803/2020\|EPI_ISL_747798\|202 | ···················· | ···················· | ···················· | ···················· | ···················· | ···················· |
| AA | 4884 hCoV-19/Denmark/DCGC-19805/2020\|EPI_ISL_747799\|202 | ···················· | ···················· | ···················· | ···················· | ···················· | ···················· |
| AA | 4885 hCoV-19/Denmark/DCGC-17465/2020\|EPI_ISL_751000\|202 | ···················· | ···················· | ···················· | ···················· | ···················· | ···················· |

AA 4886 hCoV-19/Denmark/DCGC-17467/2020|EPI_ISL_751001|202 ···················· ···················· ···················· ···················· ···················· ····················
AA 4887 hCoV-19/Denmark/DCGC-17468/2020|EPI_ISL_751002|202 ···················· ···················· ···················· ···················· ···················· ····················
AA 4888 hCoV-19/Denmark/DCGC-17469/2020|EPI_ISL_751003|202 ···················· ···················· ···················· ···················· ···················· ····················
AA 4889 hCoV-19/Denmark/DCGC-17472/2020|EPI_ISL_751004|202 ···················· ···················· ···················· ···················· ···················· ····················
AA 4890 hCoV-19/Denmark/DCGC-17474/2020|EPI_ISL_751005|202 ···················· ···················· ···················· ···················· ···················· ····················
AA 4891 hCoV-19/Denmark/DCGC-17475/2020|EPI_ISL_751006|202 ···················· ···················· ···················· ···················· ···················· ····················
AA 4892 hCoV-19/Denmark/DCGC-17476/2020|EPI_ISL_751007|202 ···················· ···················· ···················· ···················· ···················· ····················
AA 4893 hCoV-19/Denmark/DCGC-17477/2020|EPI_ISL_751008|202 ···················· ···················· ···················· ···················· ···················· ····················
AA 4894 hCoV-19/Denmark/DCGC-17478/2020|EPI_ISL_751009|202 ···················· ···················· ···················· ···················· ···················· ····················
AA 4895 hCoV-19/Denmark/DCGC-17479/2020|EPI_ISL_751010|202 ···················· ···················· ···················· ···················· ···················· ····················
AA 4896 hCoV-19/Uruguay/TYT-M136/2020|EPI_ISL_751011|2020- ···················· ···················· ···················· ···················· ···················· ····················
AA 4897 hCoV-19/Denmark/DCGC-17480/2020|EPI_ISL_751012|202 ···················· ···················· ···················· ···················· ···················· ····················
AA 4898 hCoV-19/Denmark/DCGC-17481/2020|EPI_ISL_751013|202 ···················· ···················· ···················· ···················· ···················· ····················
AA 4899 hCoV-19/Denmark/DCGC-17482/2020|EPI_ISL_751014|202 ···················· ···················· ···················· ···················· ···················· ····················
AA 4900 hCoV-19/Denmark/DCGC-17483/2020|EPI_ISL_751015|202 ···················· ···················· ···················· ···················· ···················· ····················
AA 4901 hCoV-19/Denmark/DCGC-17484/2020|EPI_ISL_751016|202 ···················· ···················· ···················· ···················· ···················· ····················
AA 4902 hCoV-19/Denmark/DCGC-17485/2020|EPI_ISL_751017|202 ···················· ···················· ···················· ···················· ···················· ····················
AA 4903 hCoV-19/Denmark/DCGC-17486/2020|EPI_ISL_751018|202 ···················· ···················· ···················· ···················· ···················· ····················
AA 4904 hCoV-19/Denmark/DCGC-17487/2020|EPI_ISL_751019|202 ···················· ···················· ···················· ···················· ···················· ····················
AA 4905 hCoV-19/Denmark/DCGC-17488/2020|EPI_ISL_751020|202 ···················· ···················· ···················· ···················· ···················· ····················
AA 4906 hCoV-19/Denmark/DCGC-17490/2020|EPI_ISL_751021|202 ···················· ···················· ···················· ···················· ···················· ····················
AA 4907 hCoV-19/Denmark/DCGC-17491/2020|EPI_ISL_751022|202 ···················· ·A·················· ···················· ···················· ···················· ··A·················
AA 4908 hCoV-19/Denmark/DCGC-17492/2020|EPI_ISL_751023|202 ···················· ···················· ···················· ···················· ···················· ····················
AA 4909 hCoV-19/Denmark/DCGC-17493/2020|EPI_ISL_751024|202 ···················· ···················· ···················· ···················· ···················· ····················
AA 4910 hCoV-19/Denmark/DCGC-17496/2020|EPI_ISL_751025|202 ···················· ···················· ···················· ···················· ···················· ····················
AA 4911 hCoV-19/Denmark/DCGC-17500/2020|EPI_ISL_751026|202 ···················· ···················· ···················· ···················· ···················· ····················
AA 4912 hCoV-19/Denmark/DCGC-17503/2020|EPI_ISL_751027|202 ···················· ···················· ···················· ···················· ···················· ····················
AA 4913 hCoV-19/Denmark/DCGC-17504/2020|EPI_ISL_751028|202 ···················· ···················· ···················· ···················· ···················· ····················
AA 4914 hCoV-19/Denmark/DCGC-17505/2020|EPI_ISL_751029|202 ···················· ·A·················· ···················· ···················· ···················· ··A·················
AA 4915 hCoV-19/Denmark/DCGC-17506/2020|EPI_ISL_751030|202 ···················· ···················· ···················· ···················· ···················· ····················
AA 4916 hCoV-19/Denmark/DCGC-17508/2020|EPI_ISL_751031|202 ···················· ···················· ···················· ···················· ···················· ····················
AA 4917 hCoV-19/Denmark/DCGC-17509/2020|EPI_ISL_751032|202 ···················· ···················· ···················· ···················· ···················· ····················
AA 4918 hCoV-19/Denmark/DCGC-17513/2020|EPI_ISL_751033|202 ···················· ···················· ···················· ···················· ···················· ····················
AA 4919 hCoV-19/Denmark/DCGC-17514/2020|EPI_ISL_751034|202 ···················· ···················· ···················· ···················· ···················· ····················
AA 4920 hCoV-19/Denmark/DCGC-17515/2020|EPI_ISL_751035|202 ···················· ···················· ···················· ···················· ···················· ····················
AA 4921 hCoV-19/Denmark/DCGC-17518/2020|EPI_ISL_751036|202 ···················· ···················· ···················· ···················· ···················· ····················
AA 4922 hCoV-19/Denmark/DCGC-17520/2020|EPI_ISL_751037|202 ···················· ···················· ···················· ···················· ···················· ····················
AA 4923 hCoV-19/USA/CA-CHLA-PLM03242140/2020|EPI_ISL_75360 ···················· ···················· ···················· ···················· ···················· ····················
AA 4924 hCoV-19/USA/CA-CHLA-PLM03369272/2020|EPI_ISL_75360 ···················· ···················· ···················· ···················· ···················· ····················
AA 4925 hCoV-19/USA/CA-CHLA-PLM58591602/2020|EPI_ISL_75360 ···················· ·A·················· ···················· ···················· ···················· ····················
AA 4926 hCoV-19/USA/CA-CHLA-PLM36106868/2020|EPI_ISL_75360 ···················· ·A·················· ···················· ···················· ···················· ····················
AA 4927 hCoV-19/USA/CA-CHLA-PLM70642018/2020|EPI_ISL_75360 ···················· ·A·················· ···················· ···················· ···················· ····················
AA 4928 hCoV-19/USA/CA-CHLA-PLM01011779/2020|EPI_ISL_75360 ···················· ·A·················· ···················· ···················· ···················· ····················
AA 4929 hCoV-19/USA/CA-CHLA-PLM81508700/2020|EPI_ISL_75360 ···················· ···················· ···················· ···················· ···················· ····················
AA 4930 hCoV-19/USA/CA-CHLA-PLM77894582/2020|EPI_ISL_75360 ···················· ···················· ···················· ···················· ···················· ····················
AA 4931 hCoV-19/USA/CA-CHLA-PLM49922269/2020|EPI_ISL_75360 ···················· ···················· ···················· ···················· ···················· ····················
AA 4932 hCoV-19/USA/CA-CHLA-PLM23092575/2020|EPI_ISL_75360 ···················· ···················· ···················· ···················· ···················· ····················
AA 4933 hCoV-19/USA/CA-CHLA-PLM41292764/2020|EPI_ISL_75361 ···················· ···················· ···················· ···················· ···················· ····················
AA 4934 hCoV-19/USA/CA-CHLA-PLM68851403/2020|EPI_ISL_75361 ···················· ···················· ···················· ···················· ···················· ····················
AA 4935 hCoV-19/USA/CA-CHLA-PLM96226608/2020|EPI_ISL_75361 ···················· ···················· ···················· ···················· ···················· ····················
AA 4936 hCoV-19/USA/CA-CHLA-PLM71497073/2020|EPI_ISL_75361 ···················· ···················· ···················· ···················· ···················· ····················
AA 4937 hCoV-19/USA/CA-CHLA-PLM71790747/2020|EPI_ISL_75361 ···················· ···················· ···················· ···················· ···················· ····················
AA 4938 hCoV-19/USA/CA-CHLA-PLM53507277/2020|EPI_ISL_75361 ···················· ···················· ···················· ···················· ···················· ····················
AA 4939 hCoV-19/USA/CA-CHLA-PLM50015204/2020|EPI_ISL_75361 ···················· ···················· ···················· ···················· ···················· ····················
AA 4940 hCoV-19/USA/CA-CHLA-PLM21825921/2020|EPI_ISL_75361 ···················· ·A·················· ···················· ···················· ···················· ····················
AA 4941 hCoV-19/USA/CA-CHLA-PLM45177859/2020|EPI_ISL_75361 ···················· ·A·················· ···················· ···················· ···················· ····················
AA 4942 hCoV-19/USA/CA-CHLA-PLM62569795/2020|EPI_ISL_75361 ···················· ··C················· ···················· ···················· ···················· ····················
AA 4943 hCoV-19/USA/CA-CHLA-PLM41556716/2020|EPI_ISL_75362 ···················· ···················· ···················· ···················· ···················· ····················
AA 4944 hCoV-19/USA/CA-CHLA-PLM80441786/2020|EPI_ISL_75365 ···················· ·A·················· ···················· ···················· ···················· ····················
AA 4945 hCoV-19/USA/CA-CHLA-PLM46887922/2020|EPI_ISL_75365 ···················· ···················· ···················· ···················· ···················· ····················
AA 4946 hCoV-19/USA/CA-CHLA-PLM09710310/2020|EPI_ISL_75365 ···················· ···················· ···················· ···················· ···················· ····················
AA 4947 hCoV-19/USA/CA-CHLA-PLM74999531/2020|EPI_ISL_75365 ···················· ···················· ···················· ···················· ···················· ····················
AA 4948 hCoV-19/USA/CA-CHLA-PLM42441520/2020|EPI_ISL_75366 ···················· ···················· ···················· ···················· ···················· ····················
AA 4949 hCoV-19/USA/CA-CHLA-PLM17644749/2020|EPI_ISL_75366 ···················· ···················· ···················· ···················· ···················· ····················
AA 4950 hCoV-19/USA/CA-CHLA-PLM01267648/2020|EPI_ISL_75366 ···················· ···················· ···················· ···················· ···················· ····················
AA 4951 hCoV-19/USA/CA-CHLA-PLM42701908/2020|EPI_ISL_75366 ···················· ···················· ···················· ···················· ···················· ····················
AA 4952 hCoV-19/USA/CA-CHLA-PLM54870960/2020|EPI_ISL_75366 ···················· ···················· ···················· ···················· ···················· ····················
AA 4953 hCoV-19/USA/CA-CHLA-PLM04997024/2020|EPI_ISL_75366 ···················· ···················· ···················· ···················· ···················· ····················
AA 4954 hCoV-19/USA/CA-CHLA-PLM94843102/2020|EPI_ISL_75366 ···················· ···················· ···················· ···················· ···················· ····················
AA 4955 hCoV-19/USA/CA-CHLA-PLM22375766/2020|EPI_ISL_75366 ···················· ···················· ···················· ···················· ···················· ····················
AA 4956 hCoV-19/USA/CA-CHLA-PLM80644511/2020|EPI_ISL_75366 ···················· ···················· ···················· ···················· ···················· ····················
AA 4957 hCoV-19/USA/CA-CHLA-PLM09699166/2020|EPI_ISL_75366 ···················· ···················· ···················· ···················· ···················· ····················
AA 4958 hCoV-19/USA/CA-CHLA-PLM41118038/2020|EPI_ISL_75367 ···················· ···················· ···················· ···················· ···················· ····················
AA 4959 hCoV-19/USA/CA-CHLA-PLM92060823/2020|EPI_ISL_75367 ···················· ···················· ···················· ···················· ···················· ····················
AA 4960 hCoV-19/USA/CA-CHLA-PLM76780180/2020|EPI_ISL_75367 ···················· ···················· ···················· ···················· ···················· ····················
AA 4961 hCoV-19/USA/CA-CHLA-PLM58754054/2020|EPI_ISL_75367 ···················· ···················· ···················· ···················· ···················· ····················
AA 4962 hCoV-19/USA/CA-CHLA-PLM39616724/2020|EPI_ISL_75367 ···················· ···················· ···················· ···················· ···················· ····················
AA 4963 hCoV-19/USA/CA-CHLA-PLM06493976/2020|EPI_ISL_75367 ···················· ···················· ···················· ···················· ···················· ····················
AA 4964 hCoV-19/USA/CA-CHLA-PLM51341617/2020|EPI_ISL_75367 ···················· ···················· ···················· ···················· ···················· ····················
AA 4965 hCoV-19/USA/CA-CHLA-PLM56652692/2020|EPI_ISL_75367 ···················· ···················· ···················· ···················· ···················· ····················

| | | | | | | | | |
|---|---|---|---|---|---|---|---|---|
| AA | 4966 | hCoV-19/USA/CA-CHLA-PLM77362941/2020\|EPI_ISL_75367 | ···················· | ···················· | ···················· | ···················· | ···················· | ···················· |
| AA | 4967 | hCoV-19/USA/CA-CHLA-PLM96511438/2020\|EPI_ISL_75367 | ···················· | ···················· | ···················· | ···················· | ···················· | ···················· |
| AA | 4968 | hCoV-19/USA/CA-CHLA-PLM99533378/2020\|EPI_ISL_75368 | ···················· | ···················· | ···················· | ···················· | ···················· | ···················· |
| AA | 4969 | hCoV-19/USA/CA-CHLA-PLM15505276/2020\|EPI_ISL_75368 | ···················· | ···················· | ···················· | ···················· | ···················· | ···················· |
| AA | 4970 | hCoV-19/USA/CA-CHLA-PLM83556703/2020\|EPI_ISL_75368 | ···················· | ···················· | ···················· | ···················· | ···················· | ···················· |
| AA | 4971 | hCoV-19/USA/CA-CHLA-PLM69425240/2020\|EPI_ISL_75368 | ···················· | ···················· | ···················· | ···················· | ···················· | ···················· |
| AA | 4972 | hCoV-19/USA/CA-CHLA-PLM57172426/2020\|EPI_ISL_75368 | ···················· | ···················· | ···················· | ···················· | ···················· | ···················· |
| AA | 4973 | hCoV-19/USA/CA-CHLA-PLM50798548/2020\|EPI_ISL_75368 | ···················· | ···················· | ···················· | ···················· | ···················· | ···················· |
| AA | 4974 | hCoV-19/USA/CA-CHLA-PLM18804118/2020\|EPI_ISL_75368 | ···················· | ···················· | ···················· | ···················· | ···················· | ···················· |
| AA | 4975 | hCoV-19/USA/CA-CHLA-PLM71552294/2020\|EPI_ISL_75368 | ···················· | ···················· | ···················· | ···················· | ···················· | ···················· |
| AA | 4976 | hCoV-19/USA/CA-CHLA-PLM60609252/2020\|EPI_ISL_75368 | ···················· | ···················· | ···················· | ···················· | ···················· | ···················· |
| AA | 4977 | hCoV-19/USA/CA-CHLA-PLM57302171/2020\|EPI_ISL_75369 | ···················· | ···················· | ···················· | ···················· | ···················· | ···················· |
| AA | 4978 | hCoV-19/USA/CA-CHLA-PLM66465316/2020\|EPI_ISL_75369 | ···················· | ···················· | ···················· | ···················· | ···················· | ···················· |
| AA | 4979 | hCoV-19/USA/CA-CHLA-PLM80048954/2020\|EPI_ISL_75369 | ···················· | ···················· | ···················· | ···················· | ···················· | ···················· |
| AA | 4980 | hCoV-19/Indonesia/KS-NIHRD-PME4901/2020\|EPI_ISL_75 | ···················· | ···················· | ···················· | ···················· | ···················· | ···················· |
| AA | 4981 | hCoV-19/USA/CA-IGI-0432/2020\|EPI_ISL_754900\|2020-0 | ···················· | ···················· | ···················· | ···················· | ···················· | ···················· |
| AA | 4982 | hCoV-19/USA/CA-IGI-0434/2020\|EPI_ISL_754901\|2020-0 | ···················· | ···················· | ···················· | ···················· | ···················· | ···················· |
| AA | 4983 | hCoV-19/Poland/Masovia-14965/2020\|EPI_ISL_754902\|2 | ···················· | ···················· | ···················· | ···················· | ···················· | ···················· |
| AA | 4984 | hCoV-19/Poland/Masovia-12865/2020\|EPI_ISL_754904\|2 | ···················· | ···················· | ···················· | ···················· | ···················· | ···················· |
| AA | 4985 | hCoV-19/Poland/Masovia-13529/2020\|EPI_ISL_754905\|2 | ···················· | ···················· | ···················· | ···················· | ···················· | ···················· |
| AA | 4986 | hCoV-19/Poland/Masovia-12866/2020\|EPI_ISL_754906\|2 | ···················· | ···················· | ···················· | ···················· | ···················· | ···················· |
| AA | 4987 | hCoV-19/Poland/Masovia-13450/2020\|EPI_ISL_754907\|2 | ···················· | ···················· | ···················· | ···················· | ···················· | ···················· |
| AA | 4988 | hCoV-19/Poland/Masovia-3342/2020\|EPI_ISL_754908\|20 | ···················· | ···················· | ···················· | ···················· | ···················· | ···················· |
| AA | 4989 | hCoV-19/Poland/Masovia-14512/2020\|EPI_ISL_754909\|2 | ···················· | ···················· | ···················· | ···················· | ···················· | ···················· |
| AA | 4990 | hCoV-19/Poland/Masovia-14790/2020\|EPI_ISL_754910\|2 | ···················· | ···················· | ···················· | ···················· | ···················· | ···················· |
| AA | 4991 | hCoV-19/Poland/Masovia-4249/2020\|EPI_ISL_754911\|20 | ···················· | ···················· | ···················· | ···················· | ···················· | ···················· |
| AA | 4992 | hCoV-19/Poland/Masovia-14376/2020\|EPI_ISL_754912\|2 | ···················· | ···················· | ···················· | ···················· | ···················· | ···················· |
| AA | 4993 | hCoV-19/Poland/Masovia-13802/2020\|EPI_ISL_754913\|2 | ···················· | ···················· | ···················· | ···················· | ···················· | ···················· |
| AA | 4994 | hCoV-19/Scotland/QEUH-C33D2F/2020\|EPI_ISL_757140\|2 | ···················· | ···················· | ···················· | ···················· | ···················· | ···················· |
| AA | 4995 | hCoV-19/Scotland/QEUH-C34641/2020\|EPI_ISL_757141\|2 | ···················· | ···················· | ···················· | ···················· | ···················· | ···················· |
| AA | 4996 | hCoV-19/Scotland/QEUH-C3367F/2020\|EPI_ISL_757142\|2 | ···················· | ···················· | ···················· | ···················· | ···················· | ···················· |
| AA | 4997 | hCoV-19/Scotland/QEUH-C3466F/2020\|EPI_ISL_757143\|2 | ···················· | ···················· | ···················· | ···················· | ···················· | ···················· |
| AA | 4998 | hCoV-19/Scotland/QEUH-C33080/2020\|EPI_ISL_757144\|2 | ···················· | ···················· | ···················· | ···················· | ···················· | ···················· |
| AA | 4999 | hCoV-19/Scotland/QEUH-C3309F/2020\|EPI_ISL_757145\|2 | ···················· | ···················· | ···················· | ···················· | ···················· | ···················· |
| AA | 5000 | hCoV-19/Scotland/QEUH-C33D5C/2020\|EPI_ISL_757146\|2 | ···················· | ···················· | ···················· | ···················· | ···················· | ···················· |
| AA | 5001 | hCoV-19/Scotland/QEUH-C3378B/2020\|EPI_ISL_757147\|2 | ···················· | ···················· | ···················· | ···················· | ···················· | ···················· |
| AA | 5002 | hCoV-19/Scotland/QEUH-C330CC/2020\|EPI_ISL_757148\|2 | ···················· | ···················· | ···················· | ···················· | ···················· | ···················· |
| AA | 5003 | hCoV-19/Scotland/QEUH-C339C1/2020\|EPI_ISL_757149\|2 | ···················· | ···················· | ···················· | ···················· | ···················· | ···················· |
| AA | 5004 | hCoV-19/Scotland/QEUH-C3477B/2020\|EPI_ISL_757150\|2 | ···················· | ···················· | ···················· | ···················· | ···················· | ···················· |
| AA | 5005 | hCoV-19/Scotland/QEUH-C339D0/2020\|EPI_ISL_757152\|2 | ···················· | ···················· | ···················· | ···················· | ···················· | ···················· |
| AA | 5006 | hCoV-19/Scotland/QEUH-C33E95/2020\|EPI_ISL_757153\|2 | ···················· | ···················· | ···················· | ···················· | ···················· | ···················· |
| AA | 5007 | hCoV-19/Scotland/QEUH-C33C8C/2020\|EPI_ISL_757154\|2 | ···················· | ···················· | ···················· | ···················· | ···················· | ···················· |
| AA | 5008 | hCoV-19/Scotland/QEUH-C3319C/2020\|EPI_ISL_757155\|2 | ···················· | ···················· | ···················· | ···················· | ···················· | ···················· |
| AA | 5009 | hCoV-19/Scotland/QEUH-C331E7/2020\|EPI_ISL_757156\|2 | ···················· | ···················· | ···················· | ···················· | ···················· | ···················· |
| AA | 5010 | hCoV-19/Scotland/QEUH-C33105/2020\|EPI_ISL_757158\|2 | ···················· | ···················· | ···················· | ···················· | ···················· | ···················· |
| AA | 5011 | hCoV-19/Scotland/QEUH-C33BF8/2020\|EPI_ISL_757159\|2 | ···················· | ···················· | ···················· | ···················· | ···················· | ···················· |
| AA | 5012 | hCoV-19/Scotland/QEUH-C33DC5/2020\|EPI_ISL_757160\|2 | ···················· | ···················· | ···················· | ···················· | ···················· | ···················· |
| AA | 5013 | hCoV-19/Scotland/QEUH-C3368E/2020\|EPI_ISL_757162\|2 | ···················· | ···················· | ···················· | ···················· | ···················· | ···················· |
| AA | 5014 | hCoV-19/Scotland/QEUH-C339A3/2020\|EPI_ISL_757163\|2 | ·····G·············· | ···················· | ···················· | ···················· | ···················· | ···················· |
| AA | 5015 | hCoV-19/Scotland/QEUH-C339EF/2020\|EPI_ISL_757164\|2 | ···················· | ···················· | ···················· | ···················· | ···················· | ···················· |
| AA | 5016 | hCoV-19/Scotland/QEUH-C3377C/2020\|EPI_ISL_757165\|2 | ···················· | ···················· | ···················· | ···················· | ······T············· | ···················· |
| AA | 5017 | hCoV-19/Scotland/QEUH-C3316F/2020\|EPI_ISL_757166\|2 | ···················· | ···················· | ···················· | ···················· | ···················· | ···················· |
| AA | 5018 | hCoV-19/Scotland/QEUH-C330F9/2020\|EPI_ISL_757167\|2 | ···················· | ···················· | ···················· | ···················· | ···················· | ···················· |
| AA | 5019 | hCoV-19/Scotland/QEUH-C33660/2020\|EPI_ISL_757168\|2 | ···················· | ···················· | ···················· | ···················· | ···················· | ···················· |
| AA | 5020 | hCoV-19/Scotland/QEUH-C33976/2020\|EPI_ISL_757169\|2 | ···················· | ···················· | ···················· | ···················· | ······T············· | ···················· |
| AA | 5021 | hCoV-19/Scotland/QEUH-C32E3C/2020\|EPI_ISL_757170\|2 | ···················· | ···················· | ···················· | ···················· | ···················· | ···················· |
| AA | 5022 | hCoV-19/Scotland/QEUH-C32E78/2020\|EPI_ISL_757171\|2 | ···················· | ···················· | ···················· | ···················· | ···················· | ···················· |
| AA | 5023 | hCoV-19/Scotland/QEUH-C33CAA/2020\|EPI_ISL_757172\|2 | ···················· | ···················· | ···················· | ···················· | ···················· | ···················· |
| AA | 5024 | hCoV-19/Scotland/QEUH-C336BB/2020\|EPI_ISL_757173\|2 | ···················· | ···················· | ···················· | ···················· | ···················· | ···················· |
| AA | 5025 | hCoV-19/Scotland/QEUH-C330DB/2020\|EPI_ISL_757174\|2 | ···················· | ···················· | ···················· | ···················· | ···················· | ···················· |
| AA | 5026 | hCoV-19/Scotland/QEUH-C33E68/2020\|EPI_ISL_757175\|2 | ···················· | ···················· | ···················· | ···················· | ···················· | ···················· |
| AA | 5027 | hCoV-19/Scotland/QEUH-C33C7D/2020\|EPI_ISL_757176\|2 | ···················· | ···················· | ···················· | ···················· | ···················· | ···················· |
| AA | 5028 | hCoV-19/Scotland/QEUH-C330AE/2020\|EPI_ISL_757177\|2 | ···················· | ···················· | ···················· | ···················· | ···················· | ···················· |
| AA | 5029 | hCoV-19/Scotland/QEUH-C330BD/2020\|EPI_ISL_757178\|2 | ···················· | ···················· | ···················· | ···················· | ···················· | ···················· |
| AA | 5030 | hCoV-19/Scotland/QEUH-C336AC/2020\|EPI_ISL_757179\|2 | ···················· | ···················· | ···················· | ···················· | ···················· | ···················· |
| AA | 5031 | hCoV-19/Scotland/QEUH-C33E0E/2020\|EPI_ISL_757180\|2 | ···················· | ···················· | ···················· | ···················· | ···················· | ···················· |
| AA | 5032 | hCoV-19/Scotland/QEUH-C33C22/2020\|EPI_ISL_757181\|2 | ···················· | ···················· | ···················· | ···················· | ···················· | ···················· |
| AA | 5033 | hCoV-19/Scotland/QEUH-C33DA7/2020\|EPI_ISL_757182\|2 | ···················· | ···················· | ···················· | ···················· | ···················· | ···················· |
| AA | 5034 | hCoV-19/Scotland/QEUH-C33958/2020\|EPI_ISL_757184\|2 | ···················· | ···················· | ···················· | ···················· | ···················· | ···················· |
| AA | 5035 | hCoV-19/Scotland/QEUH-C33985/2020\|EPI_ISL_757186\|2 | ···················· | ···················· | ···················· | ···················· | ······T············· | ···················· |
| AA | 5036 | hCoV-19/Scotland/QEUH-BA0D4C/2020\|EPI_ISL_757187\|2 | ···················· | ···················· | ···················· | ···················· | ···················· | ···················· |
| AA | 5037 | hCoV-19/Scotland/QEUH-BA1A62/2020\|EPI_ISL_757188\|2 | ···················· | ···················· | ···················· | ···················· | ···················· | ···················· |
| AA | 5038 | hCoV-19/Scotland/QEUH-BA0A27/2020\|EPI_ISL_757189\|2 | ···················· | ···················· | ···················· | ···················· | ···················· | ···················· |
| AA | 5039 | hCoV-19/Scotland/QEUH-BA0DD3/2020\|EPI_ISL_757190\|2 | ···················· | ···················· | ···················· | ···················· | ···················· | ···················· |
| AA | 5040 | hCoV-19/Scotland/QEUH-B9FF4A/2020\|EPI_ISL_757192\|2 | ···················· | ···················· | ···················· | ···················· | ···················· | ···················· |
| AA | 5041 | hCoV-19/Scotland/QEUH-B9FEC5/2020\|EPI_ISL_757193\|2 | ···················· | ···················· | ···················· | ···················· | ···················· | ···················· |
| AA | 5042 | hCoV-19/Scotland/QEUH-BA0FFB/2020\|EPI_ISL_757194\|2 | ···················· | ···················· | ···················· | ···················· | ···················· | ···················· |
| AA | 5043 | hCoV-19/Scotland/QEUH-BA165F/2020\|EPI_ISL_757195\|2 | ···················· | ···················· | ···················· | ···················· | ···················· | ···················· |
| AA | 5044 | hCoV-19/Scotland/QEUH-BA19ED/2020\|EPI_ISL_757196\|2 | ···················· | ···················· | ···················· | ···················· | ···················· | ···················· |
| AA | 5045 | hCoV-19/Scotland/QEUH-B9E646/2020\|EPI_ISL_757197\|2 | ···················· | ···················· | ···················· | ···················· | ···················· | ···················· |

| | | | | | | | |
|---|---|---|---|---|---|---|---|
| AA | 5046 hCoV-19/Scotland/QEUH-B9E655/2020\|EPI_ISL_757198\|2 | ···················· | ···················· | ····················· | ···················· | ···················· | ···················· |
| AA | 5047 hCoV-19/Denmark/DCGC-21850/2020\|EPI_ISL_758404\|202 | ···················· | ···················· | ····················· | ···················· | ···················· | ···················· |
| AA | 5048 hCoV-19/Denmark/DCGC-21851/2020\|EPI_ISL_758405\|202 | ···················· | ···················· | ····················· | ···················· | ···················· | ···················· |
| AA | 5049 hCoV-19/Denmark/DCGC-21855/2020\|EPI_ISL_758408\|202 | ···················· | ···················· | ····················· | ···················· | ···················· | ···················· |
| AA | 5050 hCoV-19/Denmark/DCGC-21856/2020\|EPI_ISL_758409\|202 | ···················· | ···················· | ····················· | ···················· | ···················· | ···················· |
| AA | 5051 hCoV-19/Denmark/DCGC-21857/2020\|EPI_ISL_758410\|202 | ···················· | ···················· | ····················· | ···················· | ···················· | ···················· |
| AA | 5052 hCoV-19/Denmark/DCGC-21858/2020\|EPI_ISL_758411\|202 | ···················· | ···················· | ····················· | ···················· | ···················· | ···················· |
| AA | 5053 hCoV-19/Denmark/DCGC-21859/2020\|EPI_ISL_758412\|202 | ···················· | ···················· | ····················· | ···················· | ···················· | ···················· |
| AA | 5054 hCoV-19/Denmark/DCGC-21860/2020\|EPI_ISL_758413\|202 | ···················· | ·A·················· | ····················· | ···················· | ···················· | ···················· |
| AA | 5055 hCoV-19/Denmark/DCGC-21861/2020\|EPI_ISL_758414\|202 | ···················· | ···················· | ····················· | ···················· | ···················· | ···················· |
| AA | 5056 hCoV-19/Denmark/DCGC-21862/2020\|EPI_ISL_758415\|202 | ···················· | ···················· | ····················· | ···················· | ···················· | ···················· |
| AA | 5057 hCoV-19/Denmark/DCGC-21863/2020\|EPI_ISL_758416\|202 | ···················· | ···················· | ····················· | ···················· | ···················· | ···················· |
| AA | 5058 hCoV-19/Denmark/DCGC-21865/2020\|EPI_ISL_758417\|202 | ···················· | ···················· | ····················· | ···················· | ···················· | ···················· |
| AA | 5059 hCoV-19/Denmark/DCGC-21867/2020\|EPI_ISL_758418\|202 | ···················· | ···················· | ····················· | ···················· | ···················· | ···················· |
| AA | 5060 hCoV-19/Denmark/DCGC-21868/2020\|EPI_ISL_758419\|202 | ···················· | ···················· | ····················· | ···················· | ···················· | ···················· |
| AA | 5061 hCoV-19/Denmark/DCGC-21869/2020\|EPI_ISL_758420\|202 | ···················· | ···················· | ····················· | ···················· | ···················· | ···················· |
| AA | 5062 hCoV-19/Denmark/DCGC-21870/2020\|EPI_ISL_758421\|202 | ···················· | ···················· | ····················· | ···················· | ···················· | ···················· |
| AA | 5063 hCoV-19/Denmark/DCGC-21871/2020\|EPI_ISL_758422\|202 | ···················· | ···················· | ····················· | ···················· | ···················· | ···················· |
| AA | 5064 hCoV-19/Denmark/DCGC-21872/2020\|EPI_ISL_758423\|202 | ···················· | ···················· | ····················· | ···················· | ···················· | ···················· |
| AA | 5065 hCoV-19/Denmark/DCGC-21874/2020\|EPI_ISL_758424\|202 | ···················· | ···················· | ····················· | ···················· | ···················· | ···················· |
| AA | 5066 hCoV-19/Denmark/DCGC-21876/2020\|EPI_ISL_758425\|202 | ···················· | ···················· | ····················· | ···················· | ···················· | ···················· |
| AA | 5067 hCoV-19/Denmark/DCGC-21878/2020\|EPI_ISL_758426\|202 | ···················· | ···················· | ····················· | ···················· | ···················· | ···················· |
| AA | 5068 hCoV-19/Denmark/DCGC-21879/2020\|EPI_ISL_758427\|202 | ···················· | ···················· | ····················· | ···················· | ···················· | ···················· |
| AA | 5069 hCoV-19/Denmark/DCGC-21881/2020\|EPI_ISL_758428\|202 | ···················· | ···················· | ····················· | ···················· | ···················· | ···················· |
| AA | 5070 hCoV-19/Denmark/DCGC-21882/2020\|EPI_ISL_758429\|202 | ···················· | ···················· | ····················· | ···················· | ···················· | ···················· |
| AA | 5071 hCoV-19/Denmark/DCGC-21946/2020\|EPI_ISL_758470\|202 | ···················· | ···················· | ····················· | ···················· | ···················· | ···················· |
| AA | 5072 hCoV-19/Denmark/DCGC-21949/2020\|EPI_ISL_758471\|202 | ···················· | ···················· | ····················· | ···················· | ···················· | ···················· |
| AA | 5073 hCoV-19/Denmark/DCGC-21950/2020\|EPI_ISL_758472\|202 | ···················· | ···················· | ····················· | ···················· | ···················· | ···················· |
| AA | 5074 hCoV-19/Denmark/DCGC-21951/2020\|EPI_ISL_758473\|202 | ···················· | ···················· | ····················· | ···················· | ···················· | ···················· |
| AA | 5075 hCoV-19/Denmark/DCGC-21952/2020\|EPI_ISL_758474\|202 | ···················· | ···················· | ····················· | ···················· | ···················· | ···················· |
| AA | 5076 hCoV-19/Denmark/DCGC-21954/2020\|EPI_ISL_758475\|202 | ···················· | ···················· | ····················· | ···················· | ···················· | ···················· |
| AA | 5077 hCoV-19/Denmark/DCGC-21956/2020\|EPI_ISL_758476\|202 | ···················· | ···················· | ····················· | ···················· | ···················· | ···················· |
| AA | 5078 hCoV-19/Denmark/DCGC-21958/2020\|EPI_ISL_758477\|202 | ···················· | ···················· | ····················· | ···················· | ···················· | ···················· |
| AA | 5079 hCoV-19/Denmark/DCGC-21960/2020\|EPI_ISL_758478\|202 | ···················· | ···················· | ····················· | ···················· | ···················· | ···················· |
| AA | 5080 hCoV-19/Denmark/DCGC-21963/2020\|EPI_ISL_758479\|202 | ···················· | ···················· | ····················· | ···················· | ···················· | ···················· |
| AA | 5081 hCoV-19/Denmark/DCGC-21964/2020\|EPI_ISL_758480\|202 | ···················· | ···················· | ····················· | ···················· | ···················· | ···················· |
| AA | 5082 hCoV-19/Denmark/DCGC-21966/2020\|EPI_ISL_758481\|202 | ···················· | ···················· | ····················· | ···················· | ···················· | ···················· |
| AA | 5083 hCoV-19/Denmark/DCGC-21968/2020\|EPI_ISL_758482\|202 | ···················· | ···················· | ····················· | ···················· | ···················· | ···················· |
| AA | 5084 hCoV-19/Denmark/DCGC-21969/2020\|EPI_ISL_758483\|202 | ···················· | ···················· | ····················· | ···················· | ···················· | ···················· |
| AA | 5085 hCoV-19/Denmark/DCGC-21972/2020\|EPI_ISL_758484\|202 | ···················· | ···················· | ····················· | ···················· | ···················· | ···················· |
| AA | 5086 hCoV-19/Denmark/DCGC-21973/2020\|EPI_ISL_758485\|202 | ···················· | ···················· | ····················· | ···················· | ···················· | ···················· |
| AA | 5087 hCoV-19/Denmark/DCGC-21975/2020\|EPI_ISL_758486\|202 | ···················· | ···················· | ····················· | ···················· | ···················· | ···················· |
| AA | 5088 hCoV-19/Denmark/DCGC-21976/2020\|EPI_ISL_758487\|202 | ···················· | ···················· | ····················· | ···················· | ···················· | ···················· |
| AA | 5089 hCoV-19/Denmark/DCGC-21979/2020\|EPI_ISL_758488\|202 | ···················· | ···················· | ····················· | ···················· | ···················· | ···················· |
| AA | 5090 hCoV-19/Denmark/DCGC-21980/2020\|EPI_ISL_758489\|202 | ···················· | ···················· | ····················· | ···················· | ···················· | ···················· |
| AA | 5091 hCoV-19/Denmark/DCGC-21982/2020\|EPI_ISL_758490\|202 | ···················· | ···················· | ····················· | ···················· | ···················· | ···················· |
| AA | 5092 hCoV-19/Denmark/DCGC-21983/2020\|EPI_ISL_758491\|202 | ···················· | ···················· | ····················· | ···················· | ···················· | ···················· |
| AA | 5093 hCoV-19/Denmark/DCGC-21984/2020\|EPI_ISL_758492\|202 | ···················· | ···················· | ····················· | ···················· | ···················· | ···················· |
| AA | 5094 hCoV-19/Denmark/DCGC-21985/2020\|EPI_ISL_758493\|202 | ···················· | ···················· | ····················· | ···················· | ···················· | ···················· |
| AA | 5095 hCoV-19/Denmark/DCGC-21987/2020\|EPI_ISL_758494\|202 | ···················· | ···················· | ····················· | ···················· | ···················· | ···················· |
| AA | 5096 hCoV-19/Denmark/DCGC-21988/2020\|EPI_ISL_758495\|202 | ···················· | ···················· | ····················· | ···················· | ···················· | ···················· |
| AA | 5097 hCoV-19/Denmark/DCGC-21989/2020\|EPI_ISL_758496\|202 | ···················· | ···················· | ····················· | ···················· | ···················· | ···················· |
| AA | 5098 hCoV-19/Denmark/DCGC-21990/2020\|EPI_ISL_758497\|202 | ···················· | ···················· | ····················· | ···················· | ···················· | ···················· |
| AA | 5099 hCoV-19/Denmark/DCGC-21991/2020\|EPI_ISL_758498\|202 | ···················· | ···················· | ····················· | ···················· | ···················· | ···················· |
| AA | 5100 hCoV-19/Denmark/DCGC-21993/2020\|EPI_ISL_758499\|202 | ···················· | ···················· | ····················· | ···················· | ···················· | ···················· |
| AA | 5101 hCoV-19/USA/WI-UW-2455/2020\|EPI_ISL_759730\|2020-12 | ···················· | ···················· | ····················· | ···················· | ···················· | ···················· |
| AA | 5105 hCoV-19/USA/WI-UW-2467/2020\|EPI_ISL_759742\|2020-12 | ···················· | ·A·················· | ····················· | ···················· | ···················· | ···················· |
| AA | 5106 hCoV-19/USA/WI-UW-2468/2020\|EPI_ISL_759743\|2020-12 | ···················· | ···················· | ····················· | ···················· | ···················· | ···················· |
| AA | 5107 hCoV-19/USA/WI-UW-2469/2020\|EPI_ISL_759744\|2020-12 | ···················· | ···················· | ····················· | ···················· | ···················· | ···················· |
| AA | 5108 hCoV-19/USA/WI-UW-2471/2020\|EPI_ISL_759745\|2020-12 | ···················· | ···················· | ····················· | ···················· | ···················· | ···················· |
| AA | 5109 hCoV-19/USA/WI-UW-2472/2020\|EPI_ISL_759746\|2020-12 | ···················· | ···················· | ····················· | ···················· | ···················· | ···················· |
| AA | 5110 hCoV-19/USA/WI-UW-2473/2020\|EPI_ISL_759747\|2020-12 | ···················· | ···················· | ····················· | ···················· | ···················· | ···················· |
| AA | 5111 hCoV-19/USA/WI-UW-2474/2020\|EPI_ISL_759748\|2020-12 | ···················· | ···················· | ····················· | ···················· | ···················· | ···················· |
| AA | 5112 hCoV-19/USA/WI-UW-2475/2020\|EPI_ISL_759749\|2020-12 | ···················· | ·A·················· | ····················· | ···················· | ···················· | ···················· |
| AA | 5113 hCoV-19/USA/WI-UW-2476/2020\|EPI_ISL_759750\|2020-12 | ···················· | ···················· | ····················· | ···················· | ···················· | ···················· |
| AA | 5114 hCoV-19/USA/WI-UW-2477/2020\|EPI_ISL_759751\|2020-12 | ···················· | ·A·················· | ····················· | ···················· | ···················· | ···················· |
| AA | 5115 hCoV-19/USA/WI-UW-2478/2020\|EPI_ISL_759752\|2020-12 | ···················· | ···················· | ····················· | ···················· | ···················· | ···················· |
| AA | 5116 hCoV-19/USA/WI-UW-2480/2020\|EPI_ISL_759754\|2020-12 | ···················· | ·A·················· | ····················· | ···················· | ···················· | ···················· |
| AA | 5117 hCoV-19/USA/WI-UW-2481/2020\|EPI_ISL_759755\|2020-12 | ···················· | ···················· | ····················· | ···················· | ···················· | ···················· |
| AA | 5118 hCoV-19/USA/WI-UW-2482/2020\|EPI_ISL_759756\|2020-12 | ···················· | ···················· | ····················· | ···················· | ···················· | ···················· |
| AA | 5119 hCoV-19/USA/WI-UW-2484/2020\|EPI_ISL_759758\|2020-12 | ···················· | ···················· | ····················· | ···················· | ···················· | ···················· |
| AA | 5120 hCoV-19/USA/WI-UW-2485/2020\|EPI_ISL_759759\|2020-12 | ···················· | ···················· | ····················· | ···················· | ···················· | ···················· |
| AA | 5121 hCoV-19/Denmark/DCGC-19806/2020\|EPI_ISL_747800\|202 | ···················· | ···················· | ····················· | ···················· | ···················· | ···················· |
| AA | 5122 hCoV-19/Denmark/DCGC-19809/2020\|EPI_ISL_747801\|202 | ···················· | ···················· | ····················· | ···················· | ···················· | ···················· |
| AA | 5123 hCoV-19/Denmark/DCGC-19810/2020\|EPI_ISL_747802\|202 | ···················· | ·A·················· | ····················· | ···················· | ···················· | ···················· |
| AA | 5124 hCoV-19/Denmark/DCGC-19811/2020\|EPI_ISL_747803\|202 | ···················· | ···················· | ····················· | ···················· | ···················· | ···················· |
| AA | 5125 hCoV-19/Denmark/DCGC-19812/2020\|EPI_ISL_747804\|202 | ···················· | ···················· | ····················· | ···················· | ···················· | ···················· |
| AA | 5126 hCoV-19/Denmark/DCGC-19813/2020\|EPI_ISL_747805\|202 | ···················· | ···················· | ····················· | ···················· | ···················· | ···················· |
| AA | 5127 hCoV-19/Denmark/DCGC-19814/2020\|EPI_ISL_747806\|202 | ···················· | ···················· | ····················· | ···················· | ···················· | ···················· |
| AA | 5128 hCoV-19/Denmark/DCGC-19817/2020\|EPI_ISL_747807\|202 | ···················· | ···················· | ····················· | ···················· | ···················· | ···················· |

| | | | | | | | |
|---|---|---|---|---|---|---|---|
| AA | 5129 hCoV-19/Denmark/DCGC-19818/2020\|EPI_ISL_747808\|202 | ···················· | ···················· | ···················· | ···················· | ···················· | ···················· |
| AA | 5130 hCoV-19/Denmark/DCGC-19820/2020\|EPI_ISL_747809\|202 | ···················· | ···················· | ···················· | ···················· | ···················· | ···················· |
| AA | 5131 hCoV-19/Denmark/DCGC-19821/2020\|EPI_ISL_747810\|202 | ···················· | ···················· | ···················· | ···················· | ···················· | ···················· |
| AA | 5132 hCoV-19/Denmark/DCGC-19822/2020\|EPI_ISL_747811\|202 | ···················· | ···················· | ···················· | ···················· | ···················· | ···················· |
| AA | 5133 hCoV-19/Denmark/DCGC-19826/2020\|EPI_ISL_747812\|202 | ···················· | ···················· | ···················· | ···················· | ···················· | ···················· |
| AA | 5134 hCoV-19/Denmark/DCGC-19827/2020\|EPI_ISL_747813\|202 | ···················· | ···················· | ···················· | ···················· | ···················· | ···················· |
| AA | 5135 hCoV-19/Denmark/DCGC-19828/2020\|EPI_ISL_747814\|202 | ···················· | ·A·················· | ···················· | ···················· | ···················· | ···················· |
| AA | 5136 hCoV-19/Denmark/DCGC-19829/2020\|EPI_ISL_747815\|202 | ···················· | ···················· | ···················· | ···················· | ···················· | ···················· |
| AA | 5137 hCoV-19/Denmark/DCGC-19830/2020\|EPI_ISL_747816\|202 | ···················· | ···················· | ···················· | ···················· | ···················· | ···················· |
| AA | 5138 hCoV-19/Denmark/DCGC-19831/2020\|EPI_ISL_747817\|202 | ···················· | ···················· | ···················· | ···················· | ···················· | ···················· |
| AA | 5139 hCoV-19/Denmark/DCGC-19832/2020\|EPI_ISL_747818\|202 | ···················· | ···················· | ···················· | ···················· | ···················· | ···················· |
| AA | 5140 hCoV-19/Denmark/DCGC-19833/2020\|EPI_ISL_747819\|202 | ···················· | ···················· | ···················· | ···················· | ···················· | ···················· |
| AA | 5141 hCoV-19/Denmark/DCGC-19834/2020\|EPI_ISL_747820\|202 | ···················· | ···················· | ···················· | ···················· | ···················· | ···················· |
| AA | 5142 hCoV-19/Denmark/DCGC-19835/2020\|EPI_ISL_747821\|202 | ···················· | ···················· | ···················· | ···················· | ···················· | ···················· |
| AA | 5143 hCoV-19/Denmark/DCGC-19838/2020\|EPI_ISL_747822\|202 | ···················· | ···················· | ···················· | ···················· | ···················· | ···················· |
| AA | 5144 hCoV-19/Denmark/DCGC-19839/2020\|EPI_ISL_747823\|202 | ···················· | ···················· | ···················· | ···················· | ···················· | ···················· |
| AA | 5145 hCoV-19/Denmark/DCGC-19840/2020\|EPI_ISL_747824\|202 | ···················· | ···················· | ···················· | ···················· | ···················· | ···················· |
| AA | 5146 hCoV-19/Denmark/DCGC-19844/2020\|EPI_ISL_747825\|202 | ···················· | ···················· | ···················· | ···················· | ···················· | ···················· |
| AA | 5147 hCoV-19/Denmark/DCGC-19845/2020\|EPI_ISL_747826\|202 | ···················· | ···················· | ···················· | ···················· | ···················· | ···················· |
| AA | 5148 hCoV-19/Denmark/DCGC-19848/2020\|EPI_ISL_747827\|202 | ···················· | ···················· | ···················· | ···················· | ···················· | ···················· |
| AA | 5149 hCoV-19/Denmark/DCGC-19850/2020\|EPI_ISL_747828\|202 | ···················· | ···················· | ···················· | ···················· | ···················· | ···················· |
| AA | 5150 hCoV-19/Denmark/DCGC-19853/2020\|EPI_ISL_747829\|202 | ···················· | ···················· | ···················· | ···················· | ···················· | ···················· |
| AA | 5151 hCoV-19/Denmark/DCGC-19854/2020\|EPI_ISL_747830\|202 | ···················· | ···················· | ···················· | ···················· | ···················· | ···················· |
| AA | 5152 hCoV-19/Denmark/DCGC-19855/2020\|EPI_ISL_747831\|202 | ···················· | ···················· | ···················· | ···················· | ···················· | ···················· |
| AA | 5153 hCoV-19/Denmark/DCGC-19857/2020\|EPI_ISL_747832\|202 | ···················· | ···················· | ···················· | ···················· | ···················· | ···················· |
| AA | 5154 hCoV-19/Denmark/DCGC-19858/2020\|EPI_ISL_747833\|202 | ···················· | ···················· | ···················· | ···················· | ···················· | ···················· |
| AA | 5155 hCoV-19/Denmark/DCGC-19859/2020\|EPI_ISL_747834\|202 | ···················· | ···················· | ···················· | ···················· | ···················· | ···················· |
| AA | 5156 hCoV-19/Denmark/DCGC-19861/2020\|EPI_ISL_747835\|202 | ···················· | ···················· | ···················· | ···················· | ···················· | ···················· |
| AA | 5157 hCoV-19/Denmark/DCGC-19863/2020\|EPI_ISL_747836\|202 | ···················· | ···················· | ···················· | ···················· | ···················· | ···················· |
| AA | 5158 hCoV-19/Denmark/DCGC-19864/2020\|EPI_ISL_747837\|202 | ···················· | ···················· | ···················· | ···················· | ···················· | ···················· |
| AA | 5159 hCoV-19/Denmark/DCGC-19867/2020\|EPI_ISL_747838\|202 | ···················· | ···················· | ···················· | ···················· | ···················· | ···················· |
| AA | 5160 hCoV-19/Denmark/DCGC-19868/2020\|EPI_ISL_747839\|202 | ···················· | ···················· | ···················· | ···················· | ···················· | ···················· |
| AA | 5161 hCoV-19/Denmark/DCGC-19869/2020\|EPI_ISL_747840\|202 | ···················· | ···················· | ···················· | ···················· | ···················· | ···················· |
| AA | 5162 hCoV-19/Denmark/DCGC-19870/2020\|EPI_ISL_747841\|202 | ···················· | ···················· | ···················· | ···················· | ···················· | ···················· |
| AA | 5163 hCoV-19/Denmark/DCGC-19871/2020\|EPI_ISL_747842\|202 | ···················· | ···················· | ···················· | ···················· | ···················· | ···················· |
| AA | 5164 hCoV-19/Denmark/DCGC-19873/2020\|EPI_ISL_747843\|202 | ···················· | ···················· | ···················· | ···················· | ···················· | ···················· |
| AA | 5165 hCoV-19/Denmark/DCGC-19874/2020\|EPI_ISL_747844\|202 | ···················· | ···················· | ···················· | ···················· | ···················· | ···················· |
| AA | 5166 hCoV-19/Denmark/DCGC-19875/2020\|EPI_ISL_747845\|202 | ···················· | ···················· | ···················· | ···················· | ···················· | ···················· |
| AA | 5167 hCoV-19/Denmark/DCGC-19876/2020\|EPI_ISL_747846\|202 | ···················· | ···················· | ···················· | ···················· | ···················· | ···················· |
| AA | 5168 hCoV-19/Denmark/DCGC-19878/2020\|EPI_ISL_747847\|202 | ···················· | ···················· | ···················· | ···················· | ···················· | ···················· |
| AA | 5169 hCoV-19/Denmark/DCGC-19879/2020\|EPI_ISL_747848\|202 | ···················· | ···················· | ···················· | ···················· | ···················· | ···················· |
| AA | 5170 hCoV-19/Denmark/DCGC-19881/2020\|EPI_ISL_747849\|202 | ···················· | ···················· | ···················· | ···················· | ···················· | ···················· |
| AA | 5171 hCoV-19/Denmark/DCGC-19882/2020\|EPI_ISL_747850\|202 | ···················· | ···················· | ···················· | ···················· | ···················· | ···················· |
| AA | 5172 hCoV-19/Denmark/DCGC-19883/2020\|EPI_ISL_747851\|202 | ···················· | ···················· | ···················· | ···················· | ···················· | ···················· |
| AA | 5173 hCoV-19/Denmark/DCGC-19886/2020\|EPI_ISL_747852\|202 | ···················· | ···················· | ···················· | ···················· | ···················· | ···················· |
| AA | 5174 hCoV-19/Denmark/DCGC-19887/2020\|EPI_ISL_747853\|202 | ···················· | ···················· | ···················· | ···················· | ···················· | ···················· |
| AA | 5175 hCoV-19/Denmark/DCGC-19891/2020\|EPI_ISL_747854\|202 | ···················· | ···················· | ···················· | ···················· | ···················· | ···················· |
| AA | 5176 hCoV-19/Denmark/DCGC-19894/2020\|EPI_ISL_747855\|202 | ···················· | ···················· | ···················· | ···················· | ···················· | ···················· |
| AA | 5177 hCoV-19/Denmark/DCGC-19896/2020\|EPI_ISL_747856\|202 | ···················· | ···················· | ···················· | ···················· | ···················· | ···················· |
| AA | 5178 hCoV-19/Denmark/DCGC-19903/2020\|EPI_ISL_747857\|202 | ···················· | ···················· | ···················· | ···················· | ···················· | ···················· |
| AA | 5179 hCoV-19/Denmark/DCGC-19904/2020\|EPI_ISL_747858\|202 | ···················· | ···················· | ···················· | ···················· | ···················· | ···················· |
| AA | 5180 hCoV-19/Denmark/DCGC-19922/2020\|EPI_ISL_747870\|202 | ···················· | ···················· | ···················· | ···················· | ···················· | ···················· |
| AA | 5181 hCoV-19/Denmark/DCGC-19926/2020\|EPI_ISL_747871\|202 | ···················· | ···················· | ···················· | ···················· | ···················· | ···················· |
| AA | 5182 hCoV-19/Denmark/DCGC-19927/2020\|EPI_ISL_747872\|202 | ···················· | ···················· | ···················· | ···················· | ···················· | ···················· |
| AA | 5183 hCoV-19/Denmark/DCGC-19928/2020\|EPI_ISL_747873\|202 | ···················· | ···················· | ···················· | ···················· | ···················· | ···················· |
| AA | 5184 hCoV-19/Denmark/DCGC-19929/2020\|EPI_ISL_747874\|202 | ···················· | ···················· | ···················· | ···················· | ···················· | ···················· |
| AA | 5185 hCoV-19/Denmark/DCGC-19930/2020\|EPI_ISL_747875\|202 | ···················· | ···················· | ···················· | ···················· | ···················· | ···················· |
| AA | 5186 hCoV-19/Denmark/DCGC-19931/2020\|EPI_ISL_747876\|202 | ···················· | ···················· | ···················· | ···················· | ···················· | ···················· |
| AA | 5187 hCoV-19/Denmark/DCGC-19937/2020\|EPI_ISL_747877\|202 | ···················· | ···················· | ···················· | ···················· | ···················· | ···················· |
| AA | 5188 hCoV-19/Denmark/DCGC-19940/2020\|EPI_ISL_747878\|202 | ···················· | ·A·················· | ···················· | ···················· | ···················· | ···················· |
| AA | 5189 hCoV-19/Denmark/DCGC-19941/2020\|EPI_ISL_747879\|202 | ···················· | ···················· | ···················· | ···················· | ···················· | ···················· |
| AA | 5190 hCoV-19/Denmark/DCGC-19943/2020\|EPI_ISL_747880\|202 | ···················· | ···················· | ···················· | ···················· | ···················· | ···················· |
| AA | 5191 hCoV-19/Denmark/DCGC-19944/2020\|EPI_ISL_747881\|202 | ···················· | ···················· | ···················· | ···················· | ···················· | ···················· |
| AA | 5192 hCoV-19/Denmark/DCGC-19946/2020\|EPI_ISL_747882\|202 | ···················· | ···················· | ···················· | ···················· | ···················· | ···················· |
| AA | 5193 hCoV-19/Denmark/DCGC-19947/2020\|EPI_ISL_747883\|202 | ···················· | ···················· | ···················· | ···················· | ···················· | ···················· |
| AA | 5194 hCoV-19/Denmark/DCGC-19948/2020\|EPI_ISL_747884\|202 | ···················· | ···················· | ···················· | ···················· | ···················· | ···················· |
| AA | 5195 hCoV-19/Denmark/DCGC-19949/2020\|EPI_ISL_747885\|202 | ···················· | ···················· | ···················· | ···················· | ···················· | ···················· |
| AA | 5196 hCoV-19/Denmark/DCGC-19950/2020\|EPI_ISL_747886\|202 | ···················· | ···················· | ···················· | ···················· | ···················· | ···················· |
| AA | 5197 hCoV-19/Denmark/DCGC-19951/2020\|EPI_ISL_747887\|202 | ···················· | ···················· | ···················· | ···················· | ···················· | ···················· |
| AA | 5198 hCoV-19/Denmark/DCGC-19952/2020\|EPI_ISL_747888\|202 | ···················· | ···················· | ···················· | ···················· | ···················· | ···················· |
| AA | 5199 hCoV-19/Denmark/DCGC-19956/2020\|EPI_ISL_747889\|202 | ···················· | ···················· | ···················· | ···················· | ···················· | ···················· |
| AA | 5200 hCoV-19/Denmark/DCGC-19960/2020\|EPI_ISL_747890\|202 | ···················· | ···················· | ···················· | ···················· | ···················· | ···················· |
| AA | 5201 hCoV-19/Denmark/DCGC-19961/2020\|EPI_ISL_747891\|202 | ···················· | ···················· | ···················· | ···················· | ···················· | ···················· |
| AA | 5202 hCoV-19/Denmark/DCGC-19962/2020\|EPI_ISL_747892\|202 | ···················· | ···················· | ···················· | ···················· | ···················· | ···················· |
| AA | 5203 hCoV-19/Denmark/DCGC-19963/2020\|EPI_ISL_747893\|202 | ···················· | ···················· | ···················· | ···················· | ···················· | ···················· |
| AA | 5204 hCoV-19/Denmark/DCGC-19964/2020\|EPI_ISL_747894\|202 | ···················· | ···················· | ···················· | ···················· | ···················· | ···················· |
| AA | 5205 hCoV-19/Denmark/DCGC-19965/2020\|EPI_ISL_747895\|202 | ···················· | ···················· | ···················· | ···················· | ···················· | ···················· |
| AA | 5206 hCoV-19/Denmark/DCGC-19966/2020\|EPI_ISL_747896\|202 | ···················· | ···················· | ···················· | ···················· | ···················· | ···················· |
| AA | 5207 hCoV-19/Denmark/DCGC-19967/2020\|EPI_ISL_747897\|202 | ···················· | ···················· | ···················· | ···················· | ···················· | ···················· |
| AA | 5208 hCoV-19/Denmark/DCGC-19968/2020\|EPI_ISL_747898\|202 | ···················· | ···················· | ···················· | ···················· | ···················· | ···················· |

AA 5209 hCoV-19/Denmark/DCGC-19970/2020|EPI_ISL_747899|202 .................... .................... .................... .................... .................... ....................
AA 5210 hCoV-19/Denmark/DCGC-17521/2020|EPI_ISL_751038|202 .................... .................... .................... .................... .................... ....................
AA 5211 hCoV-19/Denmark/DCGC-17522/2020|EPI_ISL_751039|202 .................... .................... .................... .................... .................... ....................
AA 5212 hCoV-19/Denmark/DCGC-17523/2020|EPI_ISL_751040|202 .................... .................... .................... .................... .................... ....................
AA 5213 hCoV-19/Denmark/DCGC-17524/2020|EPI_ISL_751041|202 .................... .................... .................... .................... .................... ....................
AA 5214 hCoV-19/Denmark/DCGC-17525/2020|EPI_ISL_751042|202 .................... .................... .................... .................... .................... ....................
AA 5215 hCoV-19/Denmark/DCGC-17526/2020|EPI_ISL_751043|202 .................... .................... .................... .................... .................... ....................
AA 5216 hCoV-19/Denmark/DCGC-17527/2020|EPI_ISL_751044|202 .................... .................... .................... .................... .................... ....................
AA 5217 hCoV-19/Denmark/DCGC-17528/2020|EPI_ISL_751045|202 .................... .................... .................... .................... .................... ....................
AA 5218 hCoV-19/Denmark/DCGC-17529/2020|EPI_ISL_751046|202 .................... .................... .................... .................... .................... ....................
AA 5219 hCoV-19/Denmark/DCGC-17530/2020|EPI_ISL_751047|202 .................... .................... .................... .................... .................... ....................
AA 5220 hCoV-19/Denmark/DCGC-17532/2020|EPI_ISL_751048|202 .................... .................... .................... .................... .................... ....................
AA 5221 hCoV-19/Denmark/DCGC-17533/2020|EPI_ISL_751049|202 .................... .................... .................... .................... .................... ....................
AA 5222 hCoV-19/Denmark/DCGC-17534/2020|EPI_ISL_751050|202 .................... .................... .................... .................... .................... ....................
AA 5223 hCoV-19/Denmark/DCGC-17536/2020|EPI_ISL_751051|202 .................... .A.................. .................... .................... .................... ..A.................
AA 5224 hCoV-19/Denmark/DCGC-17537/2020|EPI_ISL_751052|202 .................... .................... .................... .................... .................... ....................
AA 5225 hCoV-19/Denmark/DCGC-17538/2020|EPI_ISL_751053|202 .................... .................... .................... .................... .................... ....................
AA 5226 hCoV-19/Denmark/DCGC-17539/2020|EPI_ISL_751054|202 .................... .................... .................... .................... .................... ....................
AA 5227 hCoV-19/Denmark/DCGC-17540/2020|EPI_ISL_751055|202 .................... .................... .................... .................... .................... ....................
AA 5228 hCoV-19/Denmark/DCGC-17541/2020|EPI_ISL_751056|202 .................... .................... .................... .................... .................... ....................
AA 5229 hCoV-19/Denmark/DCGC-17542/2020|EPI_ISL_751057|202 .................... .................... .................... .................... .................... ....................
AA 5230 hCoV-19/Denmark/DCGC-17544/2020|EPI_ISL_751058|202 .................... .................... .................... .................... .................... ....................
AA 5231 hCoV-19/Denmark/DCGC-17545/2020|EPI_ISL_751059|202 .................... .................... .................... .................... .................... ....................
AA 5232 hCoV-19/Denmark/DCGC-17547/2020|EPI_ISL_751060|202 .................... .................... .................... .................... .................... ....................
AA 5233 hCoV-19/Denmark/DCGC-17548/2020|EPI_ISL_751061|202 .................... .................... .................... .................... .................... ....................
AA 5234 hCoV-19/Denmark/DCGC-17550/2020|EPI_ISL_751062|202 .................... .................... .................... .................... .................... ....................
AA 5235 hCoV-19/Denmark/DCGC-17552/2020|EPI_ISL_751063|202 .................... .................... .................... .................... .................... ....................
AA 5236 hCoV-19/Denmark/DCGC-17553/2020|EPI_ISL_751064|202 .................... .................... .................... .................... .................... ....................
AA 5237 hCoV-19/Denmark/DCGC-17554/2020|EPI_ISL_751065|202 .................... .................... .................... .................... .................... ....................
AA 5238 hCoV-19/Denmark/DCGC-17556/2020|EPI_ISL_751066|202 .................... .................... .................... .................... .................... ....................
AA 5239 hCoV-19/Denmark/DCGC-17557/2020|EPI_ISL_751067|202 .................... .................... .................... .................... .................... ....................
AA 5240 hCoV-19/Denmark/DCGC-17558/2020|EPI_ISL_751068|202 .................... .................... .................... .................... .................... ....................
AA 5241 hCoV-19/Denmark/DCGC-17559/2020|EPI_ISL_751069|202 .................... .................... .................... .................... .................... ....................
AA 5242 hCoV-19/Denmark/DCGC-17560/2020|EPI_ISL_751070|202 .................... .................... .................... .................... .................... ....................
AA 5243 hCoV-19/Denmark/DCGC-17561/2020|EPI_ISL_751071|202 .................... .................... .................... .................... .................... ....................
AA 5244 hCoV-19/Denmark/DCGC-17564/2020|EPI_ISL_751072|202 .................... .................... .................... .................... .................... ....................
AA 5245 hCoV-19/Denmark/DCGC-17565/2020|EPI_ISL_751073|202 .................... .A.................. .................... .................... .................... ....................
AA 5246 hCoV-19/Denmark/DCGC-17568/2020|EPI_ISL_751074|202 .................... .................... ..A................. .................... .................... ....................
AA 5247 hCoV-19/Denmark/DCGC-17570/2020|EPI_ISL_751075|202 .................... .................... .................... .................... .................... ....................
AA 5248 hCoV-19/Denmark/DCGC-17571/2020|EPI_ISL_751076|202 .................... .................... .................... .................... .................... ....................
AA 5249 hCoV-19/Denmark/DCGC-17575/2020|EPI_ISL_751077|202 .................... .................... .................... .................... .................... ....................
AA 5250 hCoV-19/Denmark/DCGC-17577/2020|EPI_ISL_751078|202 .................... .................... .................... .................... .................... ....................
AA 5251 hCoV-19/Denmark/DCGC-17578/2020|EPI_ISL_751079|202 .................... .................... .................... .................... .................... ....................
AA 5252 hCoV-19/Denmark/DCGC-17579/2020|EPI_ISL_751080|202 .................... .................... .................... .................... .................... ....................
AA 5253 hCoV-19/Denmark/DCGC-17580/2020|EPI_ISL_751081|202 .................... .................... .................... .................... .................... ....................
AA 5254 hCoV-19/Denmark/DCGC-17581/2020|EPI_ISL_751082|202 .................... .................... .................... .................... .................... ....................
AA 5255 hCoV-19/Denmark/DCGC-17583/2020|EPI_ISL_751083|202 .................... .................... .................... .................... .................... ....................
AA 5256 hCoV-19/Denmark/DCGC-17584/2020|EPI_ISL_751084|202 .................... .................... .................... .................... .................... ....................
AA 5257 hCoV-19/Denmark/DCGC-17585/2020|EPI_ISL_751085|202 .................... .................... .................... .................... .................... ....................
AA 5258 hCoV-19/Denmark/DCGC-17586/2020|EPI_ISL_751086|202 .................... .................... .................... .................... .................... ....................
AA 5259 hCoV-19/Denmark/DCGC-17587/2020|EPI_ISL_751087|202 .................... .................... .................... .................... .................... ....................
AA 5260 hCoV-19/Denmark/DCGC-17588/2020|EPI_ISL_751088|202 .................... .................... .................... .................... .................... ....................
AA 5261 hCoV-19/Denmark/DCGC-17589/2020|EPI_ISL_751089|202 .................... .................... .................... .................... .................... ....................
AA 5262 hCoV-19/Denmark/DCGC-17590/2020|EPI_ISL_751090|202 .................... .................... .................... .................... .................... ....................
AA 5263 hCoV-19/Denmark/DCGC-17592/2020|EPI_ISL_751091|202 .................... .................... .................... .................... .................... ....................
AA 5264 hCoV-19/Denmark/DCGC-17593/2020|EPI_ISL_751092|202 .................... .................... .................... .................... .................... ....................
AA 5265 hCoV-19/Denmark/DCGC-17594/2020|EPI_ISL_751093|202 .................... .................... .................... .................... .................... ....................
AA 5266 hCoV-19/Denmark/DCGC-17597/2020|EPI_ISL_751094|202 .................... .................... .................... .................... .................... ....................
AA 5267 hCoV-19/Denmark/DCGC-17598/2020|EPI_ISL_751095|202 .................... .................... .................... .................... .................... ....................
AA 5268 hCoV-19/Denmark/DCGC-17599/2020|EPI_ISL_751096|202 .................... .................... .................... .................... .................... ....................
AA 5269 hCoV-19/Denmark/DCGC-17600/2020|EPI_ISL_751097|202 .................... .................... .................... .................... .................... ....................
AA 5270 hCoV-19/Denmark/DCGC-17603/2020|EPI_ISL_751098|202 .................... .................... .................... .................... .................... ....................
AA 5271 hCoV-19/Denmark/DCGC-17604/2020|EPI_ISL_751099|202 .................... .................... .................... .................... .................... ....................
AA 5272 hCoV-19/USA/CA-CHLA-PLM89077418/2020|EPI_ISL_75362 .................... .................... .................... .................... .................... ....................
AA 5273 hCoV-19/USA/CA-CHLA-PLM43812599/2020|EPI_ISL_75362 .................... ..C................. .................... .................... .................... ....................
AA 5274 hCoV-19/USA/CA-CHLA-PLM67203110/2020|EPI_ISL_75362 .................... .A.................. .................... .................... .................... ....................
AA 5275 hCoV-19/USA/CA-CHLA-PLM09774868/2020|EPI_ISL_75362 .................... .................... .................... .................... .................... ....................
AA 5276 hCoV-19/USA/CA-CHLA-PLM40610335/2020|EPI_ISL_75362 .................... .................... .................... .................... .................... ....................
AA 5277 hCoV-19/USA/CA-CHLA-PLM63495917/2020|EPI_ISL_75362 .................... .................... .................... .................... .................... ....................
AA 5278 hCoV-19/USA/CA-CHLA-PLM45771852/2020|EPI_ISL_75362 .................... .................... .................... .................... .................... ....................
AA 5279 hCoV-19/USA/CA-CHLA-PLM87779966/2020|EPI_ISL_75362 .................... .................... .................... .................... .................... ....................
AA 5280 hCoV-19/USA/CA-CHLA-PLM33162258/2020|EPI_ISL_75362 .................... .................... .................... .................... .................... ....................
AA 5281 hCoV-19/USA/CA-CHLA-PLM08108247/2020|EPI_ISL_75363 .................... .................... .................... .................... .................... ....................
AA 5282 hCoV-19/USA/CA-CHLA-PLM86544901/2020|EPI_ISL_75363 .................... .................... .................... .................... .................... ....................
AA 5283 hCoV-19/USA/CA-CHLA-PLM76523568/2020|EPI_ISL_75363 .................... .A.................. .................... .................... .................... ....................
AA 5284 hCoV-19/USA/CA-CHLA-PLM15682724/2020|EPI_ISL_75363 .................... .A.................. .................... .................... .................... ....................
AA 5285 hCoV-19/USA/CA-CHLA-PLM20045761/2020|EPI_ISL_75363 .................... .................... .................... .................... .................... ....................
AA 5286 hCoV-19/USA/CA-CHLA-PLM56116031/2020|EPI_ISL_75363 .................... .................... .................... .................... .................... ....................
AA 5287 hCoV-19/USA/CA-CHLA-PLM41825337/2020|EPI_ISL_75363 .................... .................... .................... .................... .................... ....................
AA 5288 hCoV-19/USA/CA-CHLA-PLM02726594/2020|EPI_ISL_75363 .................... .................... .................... .................... .................... ....................

| | | | | | | | |
|---|---|---|---|---|---|---|---|
| AA | 5289 hCoV-19/USA/CA-CHLA-PLM20536454/2020\|EPI_ISL_75363 | ···················· | ···················· | ···················· | ···················· | ···················· | ···················· |
| AA | 5290 hCoV-19/USA/CA-CHLA-PLM71317172/2020\|EPI_ISL_75363 | ···················· | ···················· | ···················· | ···················· | ···················· | ···················· |
| AA | 5291 hCoV-19/USA/CA-CHLA-PLM16207422/2020\|EPI_ISL_75364 | ···················· | ·A·················· | ···················· | ···················· | ···················· | ···················· |
| AA | 5292 hCoV-19/USA/CA-CHLA-PLM08765144/2020\|EPI_ISL_75364 | ···················· | ·A·················· | ···················· | ···················· | ···················· | ···················· |
| AA | 5293 hCoV-19/USA/CA-CHLA-PLM50806022/2020\|EPI_ISL_75364 | ···················· | ···················· | ···················· | ···················· | ···················· | ···················· |
| AA | 5294 hCoV-19/USA/CA-CHLA-PLM75302025/2020\|EPI_ISL_75364 | ···················· | ···················· | ···················· | ···················· | ···················· | ···················· |
| AA | 5295 hCoV-19/USA/CA-CHLA-PLM56707082/2020\|EPI_ISL_75364 | ···················· | ···················· | ···················· | ···················· | ···················· | ···················· |
| AA | 5296 hCoV-19/USA/CA-CHLA-PLM40483831/2020\|EPI_ISL_75364 | ···················· | ···················· | ···················· | ···················· | ···················· | ···················· |
| AA | 5297 hCoV-19/USA/CA-CHLA-PLM16150803/2020\|EPI_ISL_75364 | ···················· | ·A·················· | ···················· | ···················· | ···················· | ···················· |
| AA | 5298 hCoV-19/USA/CA-CHLA-PLM90356598/2020\|EPI_ISL_75364 | ···················· | ·A·················· | ···················· | ···················· | ···················· | ···················· |
| AA | 5299 hCoV-19/USA/CA-CHLA-PLM63867867/2020\|EPI_ISL_75364 | ···················· | ·A·················· | ···················· | ···················· | ···················· | ···················· |
| AA | 5300 hCoV-19/USA/CA-CHLA-PLM66857248/2020\|EPI_ISL_75364 | ···················· | ·A·················· | ···················· | ···················· | ···················· | ···················· |
| AA | 5301 hCoV-19/USA/CA-CHLA-PLM97776917/2020\|EPI_ISL_75365 | ···················· | ···················· | ···················· | ···················· | ···················· | ···················· |
| AA | 5302 hCoV-19/USA/CA-CHLA-PLM51328819/2020\|EPI_ISL_75365 | ···················· | ·A·················· | ···················· | ···················· | ···················· | ···················· |
| AA | 5303 hCoV-19/USA/CA-CHLA-PLM98478969/2020\|EPI_ISL_75365 | ···················· | ···················· | ···················· | ···················· | ···················· | ···················· |
| AA | 5304 hCoV-19/USA/CA-CHLA-PLM92830927/2020\|EPI_ISL_75365 | ···················· | ·A·················· | ···················· | ···················· | ···················· | ···················· |
| AA | 5305 hCoV-19/USA/CA-CHLA-PLM04835647/2020\|EPI_ISL_75365 | ···················· | ···················· | ···················· | ···················· | ···················· | ···················· |
| AA | 5306 hCoV-19/USA/CA-CHLA-PLM75910129/2020\|EPI_ISL_75365 | ···················· | ·A·················· | ···················· | ···················· | ···················· | ···················· |
| AA | 5307 hCoV-19/Scotland/QEUH-BA1464/2020\|EPI_ISL_757221\|2 | ···················· | ···················· | ···················· | ···················· | ···················· | ···················· |
| AA | 5308 hCoV-19/Scotland/QEUH-B9E628/2020\|EPI_ISL_757222\|2 | ···················· | ···················· | ···················· | ···················· | ···················· | ···················· |
| AA | 5309 hCoV-19/Scotland/QEUH-B9E682/2020\|EPI_ISL_757224\|2 | ···················· | ···················· | ···················· | ···················· | ···················· | ···················· |
| AA | 5310 hCoV-19/Scotland/QEUH-C30E98/2020\|EPI_ISL_757225\|2 | ···················· | ···················· | ···················· | ···················· | ···················· | ···················· |
| AA | 5311 hCoV-19/Scotland/QEUH-C31750/2020\|EPI_ISL_757226\|2 | ···················· | ···················· | ···················· | ···················· | ···················· | ···················· |
| AA | 5312 hCoV-19/Scotland/QEUH-C3187B/2020\|EPI_ISL_757228\|2 | ···················· | ···················· | ···················· | ···················· | ···················· | ···················· |
| AA | 5313 hCoV-19/Scotland/QEUH-C318C6/2020\|EPI_ISL_757229\|2 | ···················· | ···················· | ···················· | ···················· | ···················· | ···················· |
| AA | 5314 hCoV-19/Scotland/QEUH-C30E3E/2020\|EPI_ISL_757230\|2 | ···················· | ···················· | ···················· | ···················· | ···················· | ···················· |
| AA | 5315 hCoV-19/Scotland/QEUH-C3185D/2020\|EPI_ISL_757231\|2 | ···················· | ·A·················· | ···················· | ···················· | ···················· | ··A················· |
| AA | 5316 hCoV-19/Scotland/QEUH-C01BA8/2020\|EPI_ISL_757232\|2 | ···················· | ···················· | ···················· | ···················· | ···················· | ···················· |
| AA | 5317 hCoV-19/Scotland/QEUH-C0149E/2020\|EPI_ISL_757233\|2 | ···················· | ···················· | ···················· | ···················· | ···················· | ···················· |
| AA | 5318 hCoV-19/Scotland/QEUH-C0113D/2020\|EPI_ISL_757235\|2 | ···················· | ···················· | ···················· | ···················· | ···················· | ···················· |
| AA | 5319 hCoV-19/Scotland/QEUH-C01D84/2020\|EPI_ISL_757236\|2 | ···················· | ···················· | ···················· | ···················· | ···················· | ···················· |
| AA | 5320 hCoV-19/Scotland/QEUH-C011C4/2020\|EPI_ISL_757237\|2 | ···················· | ···················· | ···················· | ···················· | ···················· | ···················· |
| AA | 5321 hCoV-19/Scotland/QEUH-C01B99/2020\|EPI_ISL_757238\|2 | ···················· | ···················· | ···················· | ···················· | ···················· | ···················· |
| AA | 5322 hCoV-19/Scotland/QEUH-C01188/2020\|EPI_ISL_757239\|2 | ···················· | ···················· | ···················· | ···················· | ···················· | ···················· |
| AA | 5323 hCoV-19/Scotland/QEUH-C01DB1/2020\|EPI_ISL_757240\|2 | ···················· | ···················· | ···················· | ···················· | ···················· | ···················· |
| AA | 5324 hCoV-19/Scotland/QEUH-C01D57/2020\|EPI_ISL_757241\|2 | ···················· | ···················· | ···················· | ···················· | ···················· | ···················· |
| AA | 5325 hCoV-19/Scotland/QEUH-C01197/2020\|EPI_ISL_757243\|2 | ···················· | ·A·················· | ···················· | ···················· | ···················· | ··A················· |
| AA | 5326 hCoV-19/Scotland/QEUH-C01D39/2020\|EPI_ISL_757244\|2 | ···················· | ···················· | ···················· | ···················· | ···················· | ···················· |
| AA | 5327 hCoV-19/Scotland/QEUH-C0114C/2020\|EPI_ISL_757246\|2 | ···················· | ···················· | ···················· | ···················· | ···················· | ···················· |
| AA | 5328 hCoV-19/Scotland/QEUH-C01DEE/2020\|EPI_ISL_757247\|2 | ···················· | ···················· | ···················· | ···················· | ···················· | ···················· |
| AA | 5329 hCoV-19/Scotland/QEUH-C01DA2/2020\|EPI_ISL_757248\|2 | ···················· | ···················· | ···················· | ···················· | ···················· | ···················· |
| AA | 5330 hCoV-19/Scotland/QEUH-C01D2A/2020\|EPI_ISL_757249\|2 | ···················· | ···················· | ···················· | ···················· | ···················· | ···················· |
| AA | 5331 hCoV-19/Denmark/DCGC-21995/2020\|EPI_ISL_758500\|202 | ···················· | ···················· | ···················· | ···················· | ···················· | ···················· |
| AA | 5332 hCoV-19/Denmark/DCGC-21997/2020\|EPI_ISL_758501\|202 | ···················· | ···················· | ···················· | ···················· | ···················· | ···················· |
| AA | 5333 hCoV-19/Denmark/DCGC-21998/2020\|EPI_ISL_758502\|202 | ···················· | ···················· | ···················· | ···················· | ···················· | ···················· |
| AA | 5334 hCoV-19/Denmark/DCGC-21999/2020\|EPI_ISL_758503\|202 | ···················· | ···················· | ···················· | ···················· | ···················· | ···················· |
| AA | 5335 hCoV-19/Denmark/DCGC-22000/2020\|EPI_ISL_758504\|202 | ···················· | ···················· | ···················· | ···················· | ···················· | ···················· |
| AA | 5336 hCoV-19/Denmark/DCGC-22001/2020\|EPI_ISL_758505\|202 | ···················· | ···················· | ···················· | ···················· | ···················· | ···················· |
| AA | 5337 hCoV-19/Denmark/DCGC-22002/2020\|EPI_ISL_758506\|202 | ···················· | ···················· | ···················· | ···················· | ···················· | ···················· |
| AA | 5338 hCoV-19/Denmark/DCGC-22096/2020\|EPI_ISL_758550\|202 | ···················· | ···················· | ···················· | ···················· | ···················· | ···················· |
| AA | 5339 hCoV-19/Denmark/DCGC-22101/2020\|EPI_ISL_758551\|202 | ···················· | ···················· | ···················· | ···················· | ···················· | ···················· |
| AA | 5340 hCoV-19/Denmark/DCGC-22103/2020\|EPI_ISL_758552\|202 | ···················· | ···················· | ···················· | ···················· | ···················· | ···················· |
| AA | 5341 hCoV-19/Denmark/DCGC-22105/2020\|EPI_ISL_758553\|202 | ···················· | ···················· | ···················· | ···················· | ···················· | ···················· |
| AA | 5342 hCoV-19/Denmark/DCGC-22107/2020\|EPI_ISL_758554\|202 | ···················· | ···················· | ···················· | ···················· | ···················· | ···················· |
| AA | 5343 hCoV-19/Denmark/DCGC-22108/2020\|EPI_ISL_758555\|202 | ···················· | ···················· | ···················· | ···················· | ···················· | ···················· |
| AA | 5344 hCoV-19/Denmark/DCGC-22109/2020\|EPI_ISL_758556\|202 | ···················· | ·A·················· | ···················· | ···················· | ···················· | ··A················· |
| AA | 5345 hCoV-19/Denmark/DCGC-22111/2020\|EPI_ISL_758557\|202 | ···················· | ···················· | ···················· | ···················· | ···················· | ···················· |
| AA | 5346 hCoV-19/Denmark/DCGC-22112/2020\|EPI_ISL_758558\|202 | ···················· | ·A·················· | ···················· | ···················· | ···················· | ··A················· |
| AA | 5347 hCoV-19/Denmark/DCGC-22113/2020\|EPI_ISL_758559\|202 | ···················· | ···················· | ···················· | ···················· | ···················· | ···················· |
| AA | 5348 hCoV-19/Denmark/DCGC-22114/2020\|EPI_ISL_758560\|202 | ···················· | ···················· | ···················· | ···················· | ···················· | ···················· |
| AA | 5349 hCoV-19/Denmark/DCGC-22115/2020\|EPI_ISL_758561\|202 | ···················· | ···················· | ···················· | ···················· | ···················· | ···················· |
| AA | 5350 hCoV-19/Denmark/DCGC-22116/2020\|EPI_ISL_758562\|202 | ···················· | ···················· | ···················· | ···················· | ···················· | ···················· |
| AA | 5351 hCoV-19/Denmark/DCGC-22118/2020\|EPI_ISL_758563\|202 | ···················· | ···················· | ···················· | ···················· | ···················· | ···················· |
| AA | 5352 hCoV-19/Denmark/DCGC-22119/2020\|EPI_ISL_758564\|202 | ···················· | ···················· | ···················· | ···················· | ···················· | ···················· |
| AA | 5353 hCoV-19/Denmark/DCGC-22120/2020\|EPI_ISL_758565\|202 | ···················· | ···················· | ···················· | ···················· | ···················· | ···················· |
| AA | 5354 hCoV-19/Denmark/DCGC-22123/2020\|EPI_ISL_758566\|202 | ···················· | ···················· | ···················· | ···················· | ···················· | ···················· |
| AA | 5355 hCoV-19/Denmark/DCGC-22124/2020\|EPI_ISL_758567\|202 | ···················· | ···················· | ···················· | ···················· | ···················· | ···················· |
| AA | 5356 hCoV-19/Denmark/DCGC-22125/2020\|EPI_ISL_758568\|202 | ···················· | ···················· | ···················· | ···················· | ···················· | ···················· |
| AA | 5357 hCoV-19/Denmark/DCGC-22126/2020\|EPI_ISL_758569\|202 | ···················· | ···················· | ···················· | ···················· | ···················· | ···················· |
| AA | 5358 hCoV-19/Denmark/DCGC-22127/2020\|EPI_ISL_758570\|202 | ···················· | ···················· | ···················· | ···················· | ···················· | ···················· |
| AA | 5359 hCoV-19/Denmark/DCGC-22128/2020\|EPI_ISL_758571\|202 | ···················· | ···················· | ···················· | ···················· | ···················· | ···················· |
| AA | 5360 hCoV-19/Denmark/DCGC-22129/2020\|EPI_ISL_758572\|202 | ···················· | ···················· | ···················· | ···················· | ···················· | ···················· |
| AA | 5361 hCoV-19/Denmark/DCGC-22130/2020\|EPI_ISL_758573\|202 | ···················· | ···················· | ···················· | ···················· | ···················· | ···················· |
| AA | 5362 hCoV-19/Denmark/DCGC-22131/2020\|EPI_ISL_758574\|202 | ···················· | ···················· | ···················· | ···················· | ···················· | ···················· |
| AA | 5363 hCoV-19/Denmark/DCGC-22132/2020\|EPI_ISL_758575\|202 | ···················· | ···················· | ···················· | ···················· | ···················· | ···················· |
| AA | 5364 hCoV-19/Denmark/DCGC-22135/2020\|EPI_ISL_758576\|202 | ···················· | ···················· | ···················· | ···················· | ···················· | ···················· |
| AA | 5365 hCoV-19/Denmark/DCGC-22136/2020\|EPI_ISL_758577\|202 | ···················· | ···················· | ···················· | ···················· | ···················· | ···················· |
| AA | 5366 hCoV-19/Denmark/DCGC-22137/2020\|EPI_ISL_758578\|202 | ···················· | ···················· | ···················· | ···················· | ···················· | ···················· |
| AA | 5367 hCoV-19/Denmark/DCGC-22139/2020\|EPI_ISL_758579\|202 | ···················· | ···················· | ···················· | ···················· | ···················· | ···················· |
| AA | 5368 hCoV-19/Finland/23SS14A7/2020\|EPI_ISL_759800\|2020- | ···················· | ···················· | ···················· | ···················· | ···················· | ···················· |

| | | | | | | | | |
|---|---|---|---|---|---|---|---|---|
| AA | 5369 | hCoV-19/Finland/23SS14G7/2020\|EPI_ISL_759801\|2020- | ···················· | ···················· | ···················· | ···················· | ···················· | ···················· |
| AA | 5370 | hCoV-19/Finland/23SS15G9/2020\|EPI_ISL_759802\|2020- | ···················· | ·A·················· | ···················· | ···················· | ···················· | ··A················· |
| AA | 5371 | hCoV-19/Finland/23SS15H5/2020\|EPI_ISL_759803\|2020- | ···················· | ·A·················· | ···················· | ···················· | ···················· | ··A················· |
| AA | 5372 | hCoV-19/Finland/23SS2E3/2020\|EPI_ISL_759804\|2020-0 | ···················· | ·A·················· | ···················· | ···················· | ···················· | ··A················· |
| AA | 5373 | hCoV-19/Finland/23SS4B11/2020\|EPI_ISL_759805\|2020- | ···················· | ···················· | ···················· | ···················· | ···················· | ···················· |
| AA | 5374 | hCoV-19/Finland/23SS4H6/2020\|EPI_ISL_759806\|2020-0 | ···················· | ···················· | ···················· | ···················· | ···················· | ···················· |
| AA | 5375 | hCoV-19/Finland/23SS5H7/2020\|EPI_ISL_759807\|2020-0 | ···················· | ·A·················· | ···················· | ···················· | ···················· | ··A················· |
| AA | 5376 | hCoV-19/Finland/23SS9F1/2020\|EPI_ISL_759808\|2020-0 | ···················· | ···················· | ···················· | ···················· | ···················· | ···················· |
| AA | 5377 | hCoV-19/Denmark/DCGC-19906/2020\|EPI_ISL_747859\|202 | ···················· | ···················· | ···················· | ···················· | ···················· | ···················· |
| AA | 5378 | hCoV-19/Denmark/DCGC-19908/2020\|EPI_ISL_747860\|202 | ···················· | ···················· | ···················· | ···················· | ···················· | ···················· |
| AA | 5379 | hCoV-19/Denmark/DCGC-19910/2020\|EPI_ISL_747861\|202 | ···················· | ···················· | ···················· | ···················· | ···················· | ···················· |
| AA | 5380 | hCoV-19/Denmark/DCGC-19912/2020\|EPI_ISL_747862\|202 | ···················· | ···················· | ···················· | ···················· | ···················· | ···················· |
| AA | 5381 | hCoV-19/Denmark/DCGC-19914/2020\|EPI_ISL_747863\|202 | ···················· | ···················· | ···················· | ···················· | ···················· | ···················· |
| AA | 5382 | hCoV-19/Denmark/DCGC-19915/2020\|EPI_ISL_747864\|202 | ···················· | ···················· | ···················· | ···················· | ···················· | ···················· |
| AA | 5383 | hCoV-19/Denmark/DCGC-19916/2020\|EPI_ISL_747865\|202 | ···················· | ···················· | ···················· | ···················· | ···················· | ···················· |
| AA | 5384 | hCoV-19/Denmark/DCGC-19917/2020\|EPI_ISL_747866\|202 | ···················· | ···················· | ···················· | ···················· | ···················· | ···················· |
| AA | 5385 | hCoV-19/Denmark/DCGC-19919/2020\|EPI_ISL_747867\|202 | ···················· | ···················· | ···················· | ···················· | ···················· | ···················· |
| AA | 5386 | hCoV-19/Denmark/DCGC-19920/2020\|EPI_ISL_747868\|202 | ···················· | ···················· | ···················· | ···················· | ···················· | ···················· |
| AA | 5387 | hCoV-19/Denmark/DCGC-19921/2020\|EPI_ISL_747869\|202 | ···················· | ···················· | ···················· | ···················· | ···················· | ···················· |
| AA | 5388 | hCoV-19/Denmark/DCGC-19985/2020\|EPI_ISL_747910\|202 | ···················· | ···················· | ···················· | ···················· | ···················· | ···················· |
| AA | 5389 | hCoV-19/Denmark/DCGC-19986/2020\|EPI_ISL_747911\|202 | ···················· | ···················· | ···················· | ···················· | ···················· | ···················· |
| AA | 5390 | hCoV-19/Denmark/DCGC-19987/2020\|EPI_ISL_747912\|202 | ···················· | ···················· | ···················· | ···················· | ···················· | ···················· |
| AA | 5391 | hCoV-19/Denmark/DCGC-19989/2020\|EPI_ISL_747913\|202 | ···················· | ···················· | ···················· | ···················· | ···················· | ···················· |
| AA | 5392 | hCoV-19/Denmark/DCGC-19990/2020\|EPI_ISL_747914\|202 | ···················· | ···················· | ···················· | ···················· | ···················· | ···················· |
| AA | 5393 | hCoV-19/Denmark/DCGC-19993/2020\|EPI_ISL_747915\|202 | ···················· | ···················· | ···················· | ···················· | ···················· | ···················· |
| AA | 5394 | hCoV-19/Denmark/DCGC-19994/2020\|EPI_ISL_747916\|202 | ···················· | ···················· | ···················· | ···················· | ···················· | ···················· |
| AA | 5395 | hCoV-19/Denmark/DCGC-19996/2020\|EPI_ISL_747917\|202 | ···················· | ···················· | ···················· | ···················· | ···················· | ···················· |
| AA | 5396 | hCoV-19/Denmark/DCGC-19997/2020\|EPI_ISL_747918\|202 | ···················· | ···················· | ···················· | ···················· | ···················· | ···················· |
| AA | 5397 | hCoV-19/Denmark/DCGC-19999/2020\|EPI_ISL_747919\|202 | ···················· | ···················· | ···················· | ···················· | ···················· | ···················· |
| AA | 5398 | hCoV-19/Denmark/DCGC-20000/2020\|EPI_ISL_747920\|202 | ···················· | ···················· | ···················· | ···················· | ···················· | ···················· |
| AA | 5399 | hCoV-19/Denmark/DCGC-20003/2020\|EPI_ISL_747921\|202 | ···················· | ···················· | ···················· | ···················· | ···················· | ···················· |
| AA | 5400 | hCoV-19/Denmark/DCGC-20004/2020\|EPI_ISL_747922\|202 | ···················· | ···················· | ···················· | ···················· | ···················· | ···················· |
| AA | 5401 | hCoV-19/Denmark/DCGC-20005/2020\|EPI_ISL_747923\|202 | ···················· | ···················· | ···················· | ···················· | ···················· | ···················· |
| AA | 5402 | hCoV-19/Denmark/DCGC-20006/2020\|EPI_ISL_747924\|202 | ···················· | ···················· | ···················· | ···················· | ···················· | ···················· |
| AA | 5403 | hCoV-19/Denmark/DCGC-20007/2020\|EPI_ISL_747925\|202 | ···················· | ···················· | ···················· | ···················· | ···················· | ···················· |
| AA | 5404 | hCoV-19/Denmark/DCGC-20008/2020\|EPI_ISL_747926\|202 | ···················· | ···················· | ···················· | ···················· | ···················· | ···················· |
| AA | 5405 | hCoV-19/Denmark/DCGC-20010/2020\|EPI_ISL_747927\|202 | ···················· | ···················· | ··A················· | ···················· | ···················· | ···················· |
| AA | 5406 | hCoV-19/Denmark/DCGC-20011/2020\|EPI_ISL_747928\|202 | ···················· | ···················· | ···················· | ···················· | ···················· | ···················· |
| AA | 5407 | hCoV-19/Denmark/DCGC-20013/2020\|EPI_ISL_747929\|202 | ···················· | ···················· | ···················· | ···················· | ···················· | ···················· |
| AA | 5408 | hCoV-19/Denmark/DCGC-20015/2020\|EPI_ISL_747930\|202 | ···················· | ···················· | ···················· | ···················· | ···················· | ···················· |
| AA | 5409 | hCoV-19/Denmark/DCGC-20016/2020\|EPI_ISL_747931\|202 | ···················· | ···················· | ···················· | ···················· | ···················· | ···················· |
| AA | 5410 | hCoV-19/Denmark/DCGC-20017/2020\|EPI_ISL_747932\|202 | ···················· | ···················· | ···················· | ···················· | ···················· | ···················· |
| AA | 5411 | hCoV-19/Denmark/DCGC-20018/2020\|EPI_ISL_747933\|202 | ···················· | ···················· | ···················· | ···················· | ···················· | ···················· |
| AA | 5412 | hCoV-19/Denmark/DCGC-20020/2020\|EPI_ISL_747934\|202 | ···················· | ···················· | ···················· | ···················· | ···················· | ···················· |
| AA | 5413 | hCoV-19/Denmark/DCGC-20021/2020\|EPI_ISL_747935\|202 | ···················· | ···················· | ···················· | ···················· | ···················· | ···················· |
| AA | 5414 | hCoV-19/Denmark/DCGC-17605/2020\|EPI_ISL_751100\|202 | ···················· | ···················· | ···················· | ···················· | ···················· | ···················· |
| AA | 5415 | hCoV-19/Denmark/DCGC-17609/2020\|EPI_ISL_751101\|202 | ···················· | ···················· | ···················· | ···················· | ···················· | ···················· |
| AA | 5416 | hCoV-19/Denmark/DCGC-17611/2020\|EPI_ISL_751102\|202 | ···················· | ···················· | ···················· | ···················· | ···················· | ···················· |
| AA | 5417 | hCoV-19/Denmark/DCGC-17612/2020\|EPI_ISL_751103\|202 | ···················· | ···················· | ···················· | ···················· | ···················· | ···················· |
| AA | 5418 | hCoV-19/Denmark/DCGC-17614/2020\|EPI_ISL_751104\|202 | ···················· | ···················· | ···················· | ···················· | ···················· | ···················· |
| AA | 5419 | hCoV-19/Denmark/DCGC-17615/2020\|EPI_ISL_751105\|202 | ···················· | ···················· | ···················· | ···················· | ···················· | ···················· |
| AA | 5420 | hCoV-19/Denmark/DCGC-17617/2020\|EPI_ISL_751106\|202 | ···················· | ···················· | ···················· | ···················· | ···················· | ···················· |
| AA | 5421 | hCoV-19/Denmark/DCGC-17618/2020\|EPI_ISL_751107\|202 | ···················· | ···················· | ···················· | ···················· | ···················· | ···················· |
| AA | 5422 | hCoV-19/Denmark/DCGC-17619/2020\|EPI_ISL_751108\|202 | ···················· | ···················· | ···················· | ···················· | ···················· | ···················· |
| AA | 5423 | hCoV-19/Denmark/DCGC-17620/2020\|EPI_ISL_751109\|202 | ···················· | ···················· | ···················· | ···················· | ···················· | ···················· |
| AA | 5424 | hCoV-19/Denmark/DCGC-17621/2020\|EPI_ISL_751110\|202 | ···················· | ···················· | ···················· | ···················· | ···················· | ···················· |
| AA | 5425 | hCoV-19/Denmark/DCGC-17625/2020\|EPI_ISL_751111\|202 | ···················· | ···················· | ···················· | ···················· | ···················· | ···················· |
| AA | 5426 | hCoV-19/Denmark/DCGC-17626/2020\|EPI_ISL_751112\|202 | ···················· | ···················· | ···················· | ···················· | ···················· | ···················· |
| AA | 5427 | hCoV-19/Denmark/DCGC-17631/2020\|EPI_ISL_751113\|202 | ···················· | ···················· | ···················· | ···················· | ···················· | ···················· |
| AA | 5428 | hCoV-19/Denmark/DCGC-17633/2020\|EPI_ISL_751114\|202 | ···················· | ···················· | ···················· | ···················· | ···················· | ···················· |
| AA | 5429 | hCoV-19/Denmark/DCGC-17635/2020\|EPI_ISL_751115\|202 | ···················· | ···················· | ···················· | ···················· | ···················· | ···················· |
| AA | 5430 | hCoV-19/Denmark/DCGC-17638/2020\|EPI_ISL_751116\|202 | ···················· | ···················· | ···················· | ···················· | ···················· | ···················· |
| AA | 5431 | hCoV-19/Denmark/DCGC-17639/2020\|EPI_ISL_751117\|202 | ···················· | ···················· | ···················· | ···················· | ···················· | ···················· |
| AA | 5432 | hCoV-19/Denmark/DCGC-17641/2020\|EPI_ISL_751118\|202 | ···················· | ···················· | ···················· | ···················· | ···················· | ···················· |
| AA | 5433 | hCoV-19/Denmark/DCGC-17644/2020\|EPI_ISL_751119\|202 | ···················· | ···················· | ···················· | ···················· | ···················· | ···················· |
| AA | 5434 | hCoV-19/Denmark/DCGC-17645/2020\|EPI_ISL_751120\|202 | ···················· | ···················· | ···················· | ···················· | ···················· | ···················· |
| AA | 5435 | hCoV-19/Denmark/DCGC-17646/2020\|EPI_ISL_751121\|202 | ···················· | ···················· | ···················· | ···················· | ···················· | ···················· |
| AA | 5436 | hCoV-19/Denmark/DCGC-17647/2020\|EPI_ISL_751122\|202 | ···················· | ···················· | ···················· | ···················· | ···················· | ···················· |
| AA | 5437 | hCoV-19/Denmark/DCGC-17648/2020\|EPI_ISL_751123\|202 | ···················· | ·············A······ | ···················· | ···················· | ···················· | ···················· |
| AA | 5438 | hCoV-19/Denmark/DCGC-17650/2020\|EPI_ISL_751124\|202 | ···················· | ···················· | ···················· | ···················· | ···················· | ···················· |
| AA | 5439 | hCoV-19/Denmark/DCGC-17652/2020\|EPI_ISL_751125\|202 | ···················· | ···················· | ···················· | ···················· | ···················· | ···················· |
| AA | 5440 | hCoV-19/Denmark/DCGC-17654/2020\|EPI_ISL_751126\|202 | ···················· | ···················· | ···················· | ···················· | ···················· | ···················· |
| AA | 5441 | hCoV-19/Denmark/DCGC-17656/2020\|EPI_ISL_751127\|202 | ···················· | ···················· | ···················· | ···················· | ···················· | ···················· |
| AA | 5442 | hCoV-19/Denmark/DCGC-17659/2020\|EPI_ISL_751128\|202 | ···················· | ···················· | ···················· | ···················· | ···················· | ···················· |
| AA | 5443 | hCoV-19/Denmark/DCGC-17660/2020\|EPI_ISL_751129\|202 | ···················· | ···················· | ···················· | ···················· | ···················· | ···················· |
| AA | 5444 | hCoV-19/Denmark/DCGC-17661/2020\|EPI_ISL_751130\|202 | ···················· | ···················· | ···················· | ···················· | ···················· | ···················· |
| AA | 5445 | hCoV-19/Denmark/DCGC-17665/2020\|EPI_ISL_751131\|202 | ···················· | ···················· | ···················· | ···················· | ···················· | ···················· |
| AA | 5446 | hCoV-19/Denmark/DCGC-17666/2020\|EPI_ISL_751132\|202 | ···················· | ···················· | ···················· | ···················· | ···················· | ···················· |
| AA | 5447 | hCoV-19/Denmark/DCGC-17667/2020\|EPI_ISL_751133\|202 | ···················· | ···················· | ···················· | ···················· | ···················· | ···················· |
| AA | 5448 | hCoV-19/Denmark/DCGC-17668/2020\|EPI_ISL_751134\|202 | ···················· | ···················· | ···················· | ···················· | ···················· | ···················· |

```
AA    5449 hCoV-19/Denmark/DCGC-17671/2020|EPI_ISL_751135|202   ····················  ····················  ·····················  ····················  ····················  ····················
AA    5450 hCoV-19/Denmark/DCGC-17672/2020|EPI_ISL_751136|202   ····················  ····················  ·····················  ····················  ····················  ····················
AA    5451 hCoV-19/Denmark/DCGC-17676/2020|EPI_ISL_751137|202   ····················  ····················  ·····················  ····················  ····················  ····················
AA    5452 hCoV-19/Denmark/DCGC-17677/2020|EPI_ISL_751138|202   ····················  ····················  ·····················  ····················  ····················  ····················
AA    5453 hCoV-19/Denmark/DCGC-17679/2020|EPI_ISL_751139|202   ····················  ····················  ·····················  ····················  ····················  ····················
AA    5454 hCoV-19/Denmark/DCGC-17681/2020|EPI_ISL_751140|202   ····················  ····················  ·····················  ····················  ····················  ····················
AA    5455 hCoV-19/Denmark/DCGC-17682/2020|EPI_ISL_751141|202   ····················  ····················  ·····················  ····················  ····················  ····················
AA    5456 hCoV-19/Denmark/DCGC-17683/2020|EPI_ISL_751142|202   ····················  ····················  ·····················  ····················  ····················  ····················
AA    5457 hCoV-19/Denmark/DCGC-17684/2020|EPI_ISL_751143|202   ····················  ····················  ·····················  ····················  ····················  ····················
AA    5458 hCoV-19/Denmark/DCGC-17685/2020|EPI_ISL_751144|202   ····················  ·A··················  ·····················  ····················  ····················  ····················
AA    5459 hCoV-19/Denmark/DCGC-17686/2020|EPI_ISL_751145|202   ····················  ····················  ·····················  ····················  ····················  ····················
AA    5460 hCoV-19/Denmark/DCGC-17689/2020|EPI_ISL_751146|202   ····················  ····················  ·····················  ····················  ····················  ····················
AA    5461 hCoV-19/Denmark/DCGC-17690/2020|EPI_ISL_751147|202   ····················  ····················  ·····················  ····················  ····················  ····················
AA    5462 hCoV-19/Denmark/DCGC-17691/2020|EPI_ISL_751148|202   ····················  ····················  ·····················  ····················  ····················  ····················
AA    5463 hCoV-19/Denmark/DCGC-17692/2020|EPI_ISL_751149|202   ····················  ····················  ·····················  ····················  ····················  ····················
AA    5464 hCoV-19/Denmark/DCGC-17693/2020|EPI_ISL_751150|202   ····················  ····················  ·····················  ····················  ····················  ····················
AA    5465 hCoV-19/Denmark/DCGC-17694/2020|EPI_ISL_751151|202   ····················  ····················  ·····················  ····················  ····················  ····················
AA    5466 hCoV-19/Denmark/DCGC-17695/2020|EPI_ISL_751152|202   ····················  ····················  ·····················  ····················  ····················  ····················
AA    5467 hCoV-19/Denmark/DCGC-17697/2020|EPI_ISL_751153|202   ····················  ····················  ·····················  ····················  ····················  ····················
AA    5468 hCoV-19/Denmark/DCGC-17699/2020|EPI_ISL_751154|202   ····················  ····················  ·····················  ····················  ····················  ····················
AA    5469 hCoV-19/Denmark/DCGC-17700/2020|EPI_ISL_751155|202   ····················  ····················  ·····················  ····················  ····················  ····················
AA    5470 hCoV-19/Denmark/DCGC-17704/2020|EPI_ISL_751156|202   ····················  ····················  ·····················  ····················  ····················  ····················
AA    5471 hCoV-19/Denmark/DCGC-17706/2020|EPI_ISL_751157|202   ····················  ····················  ·····················  ····················  ····················  ····················
AA    5472 hCoV-19/Denmark/DCGC-17708/2020|EPI_ISL_751158|202   ····················  ····················  ·····················  ····················  ····················  ····················
AA    5473 hCoV-19/Bangladesh/SUST-GEB-Sunamganj-3/2020|EPI_I    ····················  ····················  ·····················  ····················  ····················  ····················
AA    5474 hCoV-19/Germany/DE-BE-ChVir-W1395-5720/2020|EPI_IS   ····················  ····················  ·····················  ····················  ····················  ····················
AA    5475 hCoV-19/Germany/DE-BE-ChVir-D1944-8293/2020|EPI_IS   ····················  ····················  ··T··················  ····················  ····················  ····················
AA    5476 hCoV-19/Germany/DE-BE-ChVir-D1929-7417/2020|EPI_IS   ····················  ····················  ·····················  ····················  ····················  ····················
AA    5477 hCoV-19/Germany/DE-BE-ChVir-D1931-8262/2020|EPI_IS   ····················  ····················  ·····················  ····················  ····················  ····················
AA    5478 hCoV-19/Germany/DE-BE-ChVir-D1933-8292/2020|EPI_IS   ····················  ····················  ·····················  ····················  ····················  ····················
AA    5479 hCoV-19/Germany/DE-BE-ChVir-D1934-8278/2020|EPI_IS   ····················  ····················  ·····················  ····················  ····················  ····················
AA    5480 hCoV-19/Germany/DE-BE-ChVir-D1936-8209/2020|EPI_IS   ····················  ····················  ·····················  ····················  ····················  ····················
AA    5481 hCoV-19/Germany/DE-BE-ChVir-D1938-8233/2020|EPI_IS   ····················  ····················  ·····················  ····················  ····················  ····················
AA    5482 hCoV-19/Germany/DE-BE-ChVir-D1939-8206/2020|EPI_IS   ····················  ····················  ·····················  ····················  ····················  ····················
AA    5483 hCoV-19/Germany/DE-BE-ChVir-D1946-8235/2020|EPI_IS   ····················  ····················  ·····················  ····················  ····················  ····················
AA    5484 hCoV-19/Germany/DE-BE-ChVir-D1955-8202/2020|EPI_IS   ····················  ····················  ·····················  ····················  ····················  ····················
AA    5485 hCoV-19/Germany/DE-BE-ChVir-D2001-8219/2020|EPI_IS   ····················  ····················  ·····················  ····················  ····················  ····················
AA    5486 hCoV-19/Germany/DE-BE-ChVir-D2009-8236/2020|EPI_IS   ····················  ····················  ·····················  ····················  ····················  ····················
AA    5487 hCoV-19/Germany/DE-BE-ChVir-D2014-8229/2020|EPI_IS   ····················  ····················  ·····················  ····················  ····················  ····················
AA    5488 hCoV-19/Germany/DE-BE-ChVir-D2029-8271/2020|EPI_IS   ····················  ····················  ·····················  ····················  ····················  ····················
AA    5489 hCoV-19/Germany/DE-BE-ChVir-D2032-8244/2020|EPI_IS   ····················  ····················  ·····················  ····················  ····················  ····················
AA    5490 hCoV-19/Germany/DE-BE-ChVir-W2118-4404/2020|EPI_IS   ····················  ····················  ·····················  ····················  ····················  ····················
AA    5491 hCoV-19/Germany/DE-NW-ChVir-D2005-8224/2020|EPI_IS   ····················  ····················  ·····················  ····················  ····················  ····················
AA    5492 hCoV-19/Germany/DE-SN-ChVir-W1313-4308/2020|EPI_IS   ····················  ····················  ·····················  ····················  ····················  ····················
AA    5493 hCoV-19/Germany/DE-SN-ChVir-W1314-4321/2020|EPI_IS   ····················  ····················  ·····················  ····················  ····················  ····················
AA    5494 hCoV-19/Germany/DE-BE-ChVir-D900-8163/2020|EPI_ISL   ····················  ····················  ·····················  ····················  ····················  ····················
AA    5495 hCoV-19/Scotland/QEUH-B9E60A/2020|EPI_ISL_757200|2   ····················  ····················  ·····················  ····················  ····················  ····················
AA    5496 hCoV-19/Scotland/QEUH-B9E5B2/2020|EPI_ISL_757201|2   ····················  ····················  ·····················  ····················  ····················  ····················
AA    5497 hCoV-19/Scotland/QEUH-BA182C/2020|EPI_ISL_757202|2   ····················  ····················  ·····················  ····················  ····················  ····················
AA    5498 hCoV-19/Scotland/QEUH-B9E5D0/2020|EPI_ISL_757204|2   ····················  ····················  ·····················  ····················  ····················  ····················
AA    5499 hCoV-19/Scotland/QEUH-BA140A/2020|EPI_ISL_757207|2   ····················  ····················  ·····················  ····················  ····················  ····················
AA    5500 hCoV-19/Scotland/QEUH-BA1437/2020|EPI_ISL_757208|2   ····················  ····················  ·····················  ····················  ····················  ····················
AA    5501 hCoV-19/Scotland/QEUH-B9E5A3/2020|EPI_ISL_757209|2   ····················  ····················  ·····················  ····················  ····················  ····················
AA    5502 hCoV-19/Scotland/QEUH-BA1473/2020|EPI_ISL_757211|2   ····················  ····················  ·····················  ····················  ····················  ····················
AA    5503 hCoV-19/Scotland/QEUH-BA1428/2020|EPI_ISL_757212|2   ····················  ····················  ·····················  ····················  ····················  ····················
AA    5504 hCoV-19/Scotland/QEUH-BA14DD/2020|EPI_ISL_757213|2   ····················  ····················  ·····················  ····················  ····················  ····················
AA    5505 hCoV-19/Scotland/QEUH-BA1419/2020|EPI_ISL_757214|2   ····················  ····················  ·····················  ····················  ····················  ····················
AA    5506 hCoV-19/Scotland/QEUH-BA26D6/2020|EPI_ISL_757217|2   ····················  ····················  ·····················  ····················  ····················  ····················
AA    5507 hCoV-19/Scotland/QEUH-BA14FB/2020|EPI_ISL_757218|2   ····················  ····················  ·····················  ····················  ····················  ····················
AA    5508 hCoV-19/Scotland/QEUH-BA14BF/2020|EPI_ISL_757219|2   ····················  ····················  ·····················  ····················  ····················  ····················
AA    5509 hCoV-19/Scotland/QEUH-C01B3F/2020|EPI_ISL_757251|2   ····················  ····················  ·····················  ····················  ····················  ····················
AA    5510 hCoV-19/Scotland/QEUH-C01DDF/2020|EPI_ISL_757252|2   ····················  ····················  ·····················  ····················  ····················  ····················
AA    5511 hCoV-19/Scotland/QEUH-C01B4E/2020|EPI_ISL_757254|2   ····················  ····················  ·····················  ····················  ····················  ····················
AA    5512 hCoV-19/Scotland/QEUH-C01470/2020|EPI_ISL_757255|2   ····················  ····················  ·····················  ····················  ····················  ····················
AA    5513 hCoV-19/Scotland/QEUH-C01B5D/2020|EPI_ISL_757256|2   ····················  ····················  ·····················  ····················  ····················  ····················
AA    5514 hCoV-19/Scotland/QEUH-C01DC0/2020|EPI_ISL_757258|2   ····················  ····················  ·····················  ····················  ····················  ····················
AA    5515 hCoV-19/Scotland/QEUH-C01D93/2020|EPI_ISL_757259|2   ····················  ····················  ·····················  ····················  ····················  ····················
AA    5516 hCoV-19/Scotland/QEUH-C01B11/2020|EPI_ISL_757260|2   ····················  ····················  ·····················  ····················  ····················  ····················
AA    5517 hCoV-19/Scotland/QEUH-C01D75/2020|EPI_ISL_757261|2   ····················  ····················  ·····················  ····················  ····················  ····················
AA    5518 hCoV-19/Scotland/QEUH-C014BC/2020|EPI_ISL_757262|2   ····················  ····················  ·····················  ····················  ····················  ····················
AA    5519 hCoV-19/Scotland/QEUH-C0115B/2020|EPI_ISL_757263|2   ····················  ····················  ·····················  ····················  ····················  ····················
AA    5520 hCoV-19/Scotland/QEUH-C01179/2020|EPI_ISL_757264|2   ····················  ····················  ·····················  ····················  ····················  ····················
AA    5521 hCoV-19/Scotland/QEUH-C01B20/2020|EPI_ISL_757265|2   ····················  ····················  ·····················  ····················  ····················  ····················
AA    5522 hCoV-19/Scotland/QEUH-C01B6C/2020|EPI_ISL_757266|2   ····················  ····················  ·····················  ····················  ····················  ····················
AA    5523 hCoV-19/Scotland/QEUH-C011B5/2020|EPI_ISL_757267|2   ····················  ····················  ·····················  ····················  ····················  ····················
AA    5524 hCoV-19/Scotland/QEUH-C01B7B/2020|EPI_ISL_757268|2   ····················  ····················  ·····················  ····················  ····················  ····················
AA    5525 hCoV-19/Scotland/QEUH-C012EF/2020|EPI_ISL_757269|2   ····················  ····················  ·····················  ····················  ····················  ····················
AA    5526 hCoV-19/Scotland/QEUH-C01D66/2020|EPI_ISL_757270|2   ····················  ····················  ·····················  ····················  ····················  ····················
AA    5527 hCoV-19/Scotland/QEUH-C014AD/2020|EPI_ISL_757271|2   ····················  ····················  ·····················  ····················  ····················  ····················
AA    5528 hCoV-19/Scotland/QEUH-C0116A/2020|EPI_ISL_757272|2   ····················  ····················  ·····················  ····················  ····················  ····················
```

| | | | | | | | |
|---|---|---|---|---|---|---|---|
| AA | 5529 hCoV-19/Scotland/QEUH-B9F85E/2020\|EPI_ISL_757273\|2 | ···················· | ···················· | ···················· | ···················· | ···················· | ···················· |
| AA | 5530 hCoV-19/Scotland/QEUH-BA41A6/2020\|EPI_ISL_757274\|2 | ···················· | ···················· | ···················· | ···················· | ···················· | ···················· |
| AA | 5531 hCoV-19/Scotland/QEUH-B9FCAD/2020\|EPI_ISL_757275\|2 | ···················· | ···················· | ···················· | ···················· | ···················· | ···················· |
| AA | 5532 hCoV-19/Scotland/QEUH-BA4249/2020\|EPI_ISL_757276\|2 | ···················· | ···················· | ···················· | ···················· | ···················· | ···················· |
| AA | 5533 hCoV-19/Scotland/QEUH-B9FBFB/2020\|EPI_ISL_757277\|2 | ···················· | ···················· | ···················· | ···················· | ···················· | ···················· |
| AA | 5534 hCoV-19/Scotland/QEUH-B9FCDA/2020\|EPI_ISL_757278\|2 | ···················· | ···················· | ···················· | ···················· | ···················· | ···················· |
| AA | 5535 hCoV-19/Scotland/QEUH-B9FCF8/2020\|EPI_ISL_757279\|2 | ···················· | ···················· | ···················· | ···················· | ···················· | ···················· |
| AA | 5536 hCoV-19/Scotland/QEUH-B9FC25/2020\|EPI_ISL_757280\|2 | ···················· | ···················· | ···················· | ···················· | ···················· | ···················· |
| AA | 5537 hCoV-19/Scotland/QEUH-B9FC9E/2020\|EPI_ISL_757281\|2 | ···················· | ···················· | ···················· | ···················· | ···················· | ···················· |
| AA | 5538 hCoV-19/Scotland/QEUH-B9FBEC/2020\|EPI_ISL_757282\|2 | ···················· | ···················· | ···················· | ···················· | ···················· | ···················· |
| AA | 5539 hCoV-19/Scotland/QEUH-B9FCCB/2020\|EPI_ISL_757283\|2 | ···················· | ···················· | ···················· | ···················· | ···················· | ···················· |
| AA | 5540 hCoV-19/Scotland/QEUH-B9FC61/2020\|EPI_ISL_757284\|2 | ···················· | ···················· | ···················· | ···················· | ···················· | ···················· |
| AA | 5541 hCoV-19/Indonesia/AC-NIHRD-C0057926/2020\|EPI_ISL_7 | ···················· | ···················· | ···················· | ···················· | ···················· | ··A················· |
| AA | 5542 hCoV-19/Finland/12MR1cG4/2020\|EPI_ISL_757288\|2020- | ···················· | ···················· | ···················· | ···················· | ···················· | ···················· |
| AA | 5543 hCoV-19/Finland/13MR4cC4/2020\|EPI_ISL_757289\|2020- | ···················· | ···················· | ···················· | ···················· | ···················· | ···················· |
| AA | 5544 hCoV-19/Finland/13MR5cB2/2020\|EPI_ISL_757290\|2020- | ···················· | ·A·················· | ···················· | ···················· | ···················· | ··A················· |
| AA | 5545 hCoV-19/Finland/14MR1cE3/2020\|EPI_ISL_757291\|2020- | ···················· | ·A·················· | ···················· | ···················· | ···················· | ··A················· |
| AA | 5546 hCoV-19/Finland/14MR1cG3/2020\|EPI_ISL_757292\|2020- | ···················· | ·A·················· | ···················· | ···················· | ···················· | ··A················· |
| AA | 5547 hCoV-19/Finland/14MR8cB5/2020\|EPI_ISL_757293\|2020- | ···················· | ···················· | ···················· | ···················· | ···················· | ···················· |
| AA | 5548 hCoV-19/Finland/14MR8cG7/2020\|EPI_ISL_757294\|2020- | ···················· | ···················· | ···················· | ···················· | ···················· | ···················· |
| AA | 5549 hCoV-19/Finland/15MR1cB4/2020\|EPI_ISL_757295\|2020- | ···················· | ···················· | ···················· | ···················· | ···················· | ···················· |
| AA | 5550 hCoV-19/Finland/16MR7cA4/2020\|EPI_ISL_757296\|2020- | ···················· | ·A·················· | ···················· | ···················· | ···················· | ··A················· |
| AA | 5551 hCoV-19/Finland/16MR7cG11/2020\|EPI_ISL_757297\|2020 | ···················· | ···················· | ···················· | ···················· | ···················· | ···················· |
| AA | 5552 hCoV-19/Finland/16MR8cH7/2020\|EPI_ISL_757298\|2020- | ···················· | ···················· | ···················· | ···················· | ···················· | ···················· |
| AA | 5553 hCoV-19/Finland/16MR9cA1/2020\|EPI_ISL_757299\|2020- | ···················· | ···················· | ···················· | ···················· | ···················· | ···················· |
| AA | 5554 hCoV-19/Denmark/DCGC-22003/2020\|EPI_ISL_758507\|202 | ···················· | ···················· | ···················· | ···················· | ···················· | ···················· |
| AA | 5555 hCoV-19/Denmark/DCGC-22005/2020\|EPI_ISL_758508\|202 | ···················· | ···················· | ···················· | ···················· | ···················· | ···················· |
| AA | 5556 hCoV-19/Denmark/DCGC-22006/2020\|EPI_ISL_758509\|202 | ···················· | ···················· | ···················· | ···················· | ···················· | ···················· |
| AA | 5557 hCoV-19/Denmark/DCGC-22007/2020\|EPI_ISL_758510\|202 | ···················· | ···················· | ···················· | ···················· | ···················· | ···················· |
| AA | 5558 hCoV-19/Denmark/DCGC-22009/2020\|EPI_ISL_758511\|202 | ···················· | ···················· | ···················· | ···················· | ···················· | ···················· |
| AA | 5559 hCoV-19/Denmark/DCGC-22015/2020\|EPI_ISL_758512\|202 | ···················· | ···················· | ···················· | ···················· | ···················· | ···················· |
| AA | 5560 hCoV-19/Denmark/DCGC-22020/2020\|EPI_ISL_758513\|202 | ···················· | ···················· | ···················· | ···················· | ···················· | ···················· |
| AA | 5561 hCoV-19/Denmark/DCGC-22021/2020\|EPI_ISL_758514\|202 | ···················· | ···················· | ···················· | ···················· | ···················· | ···················· |
| AA | 5562 hCoV-19/Denmark/DCGC-22023/2020\|EPI_ISL_758515\|202 | ···················· | ···················· | ···················· | ···················· | ···················· | ···················· |
| AA | 5563 hCoV-19/Denmark/DCGC-22024/2020\|EPI_ISL_758516\|202 | ···················· | ···················· | ···················· | ···················· | ···················· | ···················· |
| AA | 5564 hCoV-19/Denmark/DCGC-22025/2020\|EPI_ISL_758517\|202 | ···················· | ···················· | ···················· | ···················· | ···················· | ···················· |
| AA | 5565 hCoV-19/Denmark/DCGC-22029/2020\|EPI_ISL_758518\|202 | ···················· | ···················· | ···················· | ···················· | ···················· | ···················· |
| AA | 5566 hCoV-19/Denmark/DCGC-22033/2020\|EPI_ISL_758519\|202 | ···················· | ···················· | ···················· | ···················· | ···················· | ···················· |
| AA | 5567 hCoV-19/Denmark/DCGC-22035/2020\|EPI_ISL_758520\|202 | ···················· | ···················· | ···················· | ···················· | ···················· | ···················· |
| AA | 5568 hCoV-19/Denmark/DCGC-22036/2020\|EPI_ISL_758521\|202 | ···················· | ···················· | ···················· | ···················· | ···················· | ···················· |
| AA | 5569 hCoV-19/Denmark/DCGC-22038/2020\|EPI_ISL_758522\|202 | ···················· | ···················· | ···················· | ···················· | ···················· | ···················· |
| AA | 5570 hCoV-19/Denmark/DCGC-22041/2020\|EPI_ISL_758523\|202 | ···················· | ···················· | ···················· | ···················· | ···················· | ···················· |
| AA | 5571 hCoV-19/Denmark/DCGC-22042/2020\|EPI_ISL_758524\|202 | ···················· | ···················· | ···················· | ···················· | ···················· | ···················· |
| AA | 5572 hCoV-19/Denmark/DCGC-22046/2020\|EPI_ISL_758525\|202 | ···················· | ···················· | ···················· | ···················· | ···················· | ···················· |
| AA | 5573 hCoV-19/Denmark/DCGC-22049/2020\|EPI_ISL_758526\|202 | ···················· | ···················· | ···················· | ···················· | ···················· | ···················· |
| AA | 5574 hCoV-19/Denmark/DCGC-22050/2020\|EPI_ISL_758527\|202 | ···················· | ···················· | ···················· | ···················· | ···················· | ···················· |
| AA | 5575 hCoV-19/Denmark/DCGC-22052/2020\|EPI_ISL_758528\|202 | ···················· | ···················· | ···················· | ···················· | ···················· | ···················· |
| AA | 5576 hCoV-19/Denmark/DCGC-22056/2020\|EPI_ISL_758529\|202 | ···················· | ···················· | ···················· | ···················· | ···················· | ···················· |
| AA | 5577 hCoV-19/Denmark/DCGC-22057/2020\|EPI_ISL_758530\|202 | ···················· | ···················· | ···················· | ···················· | ···················· | ···················· |
| AA | 5578 hCoV-19/Denmark/DCGC-22061/2020\|EPI_ISL_758531\|202 | ···················· | ···················· | ···················· | ···················· | ···················· | ···················· |
| AA | 5579 hCoV-19/Denmark/DCGC-22062/2020\|EPI_ISL_758532\|202 | ···················· | ···················· | ···················· | ···················· | ···················· | ···················· |
| AA | 5580 hCoV-19/Denmark/DCGC-22064/2020\|EPI_ISL_758533\|202 | ···················· | ···················· | ···················· | ···················· | ···················· | ···················· |
| AA | 5581 hCoV-19/Denmark/DCGC-22066/2020\|EPI_ISL_758534\|202 | ···················· | ···················· | ···················· | ···················· | ···················· | ···················· |
| AA | 5582 hCoV-19/Denmark/DCGC-22069/2020\|EPI_ISL_758535\|202 | ···················· | ···················· | ···················· | ···················· | ···················· | ···················· |
| AA | 5583 hCoV-19/Denmark/DCGC-22070/2020\|EPI_ISL_758536\|202 | ···················· | ···················· | ···················· | ···················· | ···················· | ···················· |
| AA | 5584 hCoV-19/Denmark/DCGC-22071/2020\|EPI_ISL_758537\|202 | ···················· | ···················· | ···················· | ···················· | ···················· | ···················· |
| AA | 5585 hCoV-19/Denmark/DCGC-22075/2020\|EPI_ISL_758538\|202 | ···················· | ···················· | ···················· | ···················· | ···················· | ···················· |
| AA | 5586 hCoV-19/Denmark/DCGC-22077/2020\|EPI_ISL_758539\|202 | ···················· | ···················· | ···················· | ···················· | ···················· | ···················· |
| AA | 5587 hCoV-19/Denmark/DCGC-22078/2020\|EPI_ISL_758540\|202 | ···················· | ···················· | ···················· | ···················· | ···················· | ···················· |
| AA | 5588 hCoV-19/Denmark/DCGC-22082/2020\|EPI_ISL_758541\|202 | ···················· | ···················· | ···················· | ···················· | ···················· | ···················· |
| AA | 5589 hCoV-19/Denmark/DCGC-22084/2020\|EPI_ISL_758542\|202 | ···················· | ···················· | ···················· | ···················· | ···················· | ···················· |
| AA | 5590 hCoV-19/Denmark/DCGC-22085/2020\|EPI_ISL_758543\|202 | ···················· | ···················· | ···················· | ···················· | ···················· | ···················· |
| AA | 5591 hCoV-19/Denmark/DCGC-22086/2020\|EPI_ISL_758544\|202 | ···················· | ···················· | ···················· | ···················· | ···················· | ···················· |
| AA | 5592 hCoV-19/Denmark/DCGC-22087/2020\|EPI_ISL_758545\|202 | ···················· | ···················· | ···················· | ···················· | ···················· | ···················· |
| AA | 5593 hCoV-19/Denmark/DCGC-22088/2020\|EPI_ISL_758546\|202 | ···················· | ···················· | ···················· | ···················· | ···················· | ···················· |
| AA | 5594 hCoV-19/Denmark/DCGC-22089/2020\|EPI_ISL_758547\|202 | ···················· | ···················· | ···················· | ···················· | ···················· | ···················· |
| AA | 5595 hCoV-19/Denmark/DCGC-22091/2020\|EPI_ISL_758548\|202 | ···················· | ···················· | ···················· | ···················· | ···················· | ···················· |
| AA | 5596 hCoV-19/Denmark/DCGC-22093/2020\|EPI_ISL_758549\|202 | ···················· | ···················· | ···················· | ···················· | ···················· | ···················· |
| AA | 5597 hCoV-19/Denmark/DCGC-22140/2020\|EPI_ISL_758580\|202 | ···················· | ···················· | ···················· | ···················· | ···················· | ···················· |
| AA | 5598 hCoV-19/Denmark/DCGC-22141/2020\|EPI_ISL_758581\|202 | ···················· | ···················· | ···················· | ···················· | ···················· | ···················· |
| AA | 5599 hCoV-19/Denmark/DCGC-22144/2020\|EPI_ISL_758582\|202 | ···················· | ···················· | ···················· | ···················· | ···················· | ···················· |
| AA | 5600 hCoV-19/Denmark/DCGC-22145/2020\|EPI_ISL_758583\|202 | ···················· | ···················· | ···················· | ···················· | ···················· | ···················· |
| AA | 5601 hCoV-19/Denmark/DCGC-22146/2020\|EPI_ISL_758584\|202 | ···················· | ···················· | ···················· | ···················· | ···················· | ···················· |
| AA | 5602 hCoV-19/Denmark/DCGC-22147/2020\|EPI_ISL_758585\|202 | ···················· | ···················· | ···················· | ···················· | ···················· | ···················· |
| AA | 5603 hCoV-19/Denmark/DCGC-22148/2020\|EPI_ISL_758586\|202 | ···················· | ···················· | ···················· | ···················· | ···················· | ···················· |
| AA | 5604 hCoV-19/Denmark/DCGC-22149/2020\|EPI_ISL_758587\|202 | ···················· | ···················· | ···················· | ···················· | ···················· | ···················· |
| AA | 5605 hCoV-19/Denmark/DCGC-22150/2020\|EPI_ISL_758588\|202 | ···················· | ···················· | ···················· | ···················· | ···················· | ···················· |
| AA | 5606 hCoV-19/Denmark/DCGC-22152/2020\|EPI_ISL_758589\|202 | ···················· | ···················· | ···················· | ···················· | ···················· | ···················· |
| AA | 5607 hCoV-19/Denmark/DCGC-22153/2020\|EPI_ISL_758590\|202 | ···················· | ···················· | ···················· | ···················· | ···················· | ···················· |
| AA | 5608 hCoV-19/Denmark/DCGC-22154/2020\|EPI_ISL_758591\|202 | ···················· | ···················· | ···················· | ···················· | ···················· | ···················· |

```
AA  5609 hCoV-19/Denmark/DCGC-22157/2020|EPI_ISL_758592|202  ···················· ···················· ···················· ···················· ···················· ····················
AA  5610 hCoV-19/Denmark/DCGC-22161/2020|EPI_ISL_758593|202  ···················· ···················· ···················· ···················· ···················· ···············A····
AA  5611 hCoV-19/Denmark/DCGC-22162/2020|EPI_ISL_758594|202  ···················· ···················· ···················· ···················· ···················· ····················
AA  5612 hCoV-19/Denmark/DCGC-22164/2020|EPI_ISL_758595|202  ···················· ···················· ···················· ···················· ···················· ····················
AA  5613 hCoV-19/Denmark/DCGC-22166/2020|EPI_ISL_758596|202  ···················· ···················· ···················· ···················· ···················· ····················
AA  5614 hCoV-19/Denmark/DCGC-22170/2020|EPI_ISL_758597|202  ···················· ···················· ···················· ···················· ···················· ····················
AA  5615 hCoV-19/Denmark/DCGC-22171/2020|EPI_ISL_758598|202  ···················· ···················· ···················· ···················· ···················· ····················
AA  5616 hCoV-19/Denmark/DCGC-22174/2020|EPI_ISL_758599|202  ···················· ···················· ···················· ···················· ···················· ····················
AA  5617 hCoV-19/Denmark/DCGC-22176/2020|EPI_ISL_758600|202  ···················· ···················· ···················· ···················· ···················· ····················
AA  5618 hCoV-19/Denmark/DCGC-22179/2020|EPI_ISL_758601|202  ···················· ···················· ···················· ···················· ···················· ····················
AA  5619 hCoV-19/Denmark/DCGC-22181/2020|EPI_ISL_758602|202  ···················· ···················· ···················· ···················· ···················· ····················
AA  5620 hCoV-19/Denmark/DCGC-22186/2020|EPI_ISL_758603|202  ···················· ···················· ···················· ···················· ···················· ····················
AA  5621 hCoV-19/Denmark/DCGC-22187/2020|EPI_ISL_758604|202  ···················· ···················· ···················· ···················· ···················· ····················
AA  5622 hCoV-19/Denmark/DCGC-22188/2020|EPI_ISL_758605|202  ···················· ···················· ···················· ···················· ···················· ····················
AA  5623 hCoV-19/Denmark/DCGC-22196/2020|EPI_ISL_758608|202  ···················· ···················· ···················· ···················· ···················· ····················
AA  5624 hCoV-19/Denmark/DCGC-22197/2020|EPI_ISL_758609|202  ···················· ···················· ···················· ···················· ···················· ····················
AA  5625 hCoV-19/USA/UN-PB-Wuhan-Hu-1/2020|EPI_ISL_759860|2  ···················· ···················· ···················· ···················· ···················· ····················
AA  5626 hCoV-19/USA/UN-PB-Wuhan-Hu-1-2/2020|EPI_ISL_759861  ···················· ···················· ···················· ···················· ···················· ····················
AA  5627 hCoV-19/USA/UN-PB-Wuhan-Hu-1-3/2020|EPI_ISL_759862  ···················· ···················· ···················· ···················· ···················· ····················
AA  5628 hCoV-19/USA/UN-PB-Wuhan-Hu-1-4/2020|EPI_ISL_759863  ···················· ···················· ···················· ···················· ···················· ····················
AA  5629 hCoV-19/USA/UN-PB-Wuhan-Hu-1-5/2020|EPI_ISL_759864  ···················· ···················· ···················· ···················· ···················· ····················
AA  5630 hCoV-19/USA/UN-PB-Wuhan-Hu-1-6/2020|EPI_ISL_759865  ···················· ···················· ···················· ···················· ···················· ····················
AA  5631 hCoV-19/India/UN-NITMA-1086/2020|EPI_ISL_759866|20  ···················· ···················· ···················· ···················· ···················· ····················
AA  5632 hCoV-19/India/UN-NITMA-1139/2020|EPI_ISL_759867|20  ···················· ···················· ···················· ···················· ···················· ····················
AA  5633 hCoV-19/Denmark/DCGC-19971/2020|EPI_ISL_747900|202  ···················· ···················· ···················· ···················· ···················· ····················
AA  5634 hCoV-19/Denmark/DCGC-19974/2020|EPI_ISL_747901|202  ···················· ···················· ···················· ···················· ···················· ····················
AA  5635 hCoV-19/Denmark/DCGC-19975/2020|EPI_ISL_747902|202  ···················· ·A·················· ···················· ···················· ···················· ····················
AA  5636 hCoV-19/Denmark/DCGC-19977/2020|EPI_ISL_747903|202  ···················· ···················· ···················· ···················· ······T············· ····················
AA  5637 hCoV-19/Denmark/DCGC-19979/2020|EPI_ISL_747904|202  ···················· ···················· ···················· ···················· ···················· ····················
AA  5638 hCoV-19/Denmark/DCGC-19980/2020|EPI_ISL_747905|202  ···················· ···················· ···················· ···················· ···················· ····················
AA  5639 hCoV-19/Denmark/DCGC-19981/2020|EPI_ISL_747906|202  ···················· ···················· ···················· ···················· ···················· ····················
AA  5640 hCoV-19/Denmark/DCGC-19982/2020|EPI_ISL_747907|202  ···················· ···················· ···················· ···················· ···················· ····················
AA  5641 hCoV-19/Denmark/DCGC-19983/2020|EPI_ISL_747908|202  ···················· ···················· ···················· ···················· ···················· ····················
AA  5642 hCoV-19/Denmark/DCGC-19984/2020|EPI_ISL_747909|202  ···················· ···················· ···················· ···················· ···················· ····················
AA  5643 hCoV-19/Denmark/DCGC-20022/2020|EPI_ISL_747936|202  ···················· ·A·················· ···················· ···················· ···················· ····················
AA  5644 hCoV-19/Denmark/DCGC-20023/2020|EPI_ISL_747937|202  ···················· ···················· ···················· ···················· ···················· ····················
AA  5645 hCoV-19/Denmark/DCGC-20024/2020|EPI_ISL_747938|202  ···················· ···················· ···················· ···················· ···················· ····················
AA  5646 hCoV-19/Denmark/DCGC-20025/2020|EPI_ISL_747939|202  ···················· ···················· ···················· ···················· ···················· ····················
AA  5647 hCoV-19/Denmark/DCGC-20027/2020|EPI_ISL_747940|202  ···················· ···················· ···················· ···················· ···················· ····················
AA  5648 hCoV-19/Denmark/DCGC-20028/2020|EPI_ISL_747941|202  ···················· ···················· ···················· ···················· ···················· ····················
AA  5649 hCoV-19/Denmark/DCGC-20032/2020|EPI_ISL_747942|202  ···················· ···················· ···················· ···················· ···················· ····················
AA  5650 hCoV-19/Denmark/DCGC-20033/2020|EPI_ISL_747943|202  ···················· ···················· ···················· ···················· ···················· ····················
AA  5651 hCoV-19/Denmark/DCGC-20034/2020|EPI_ISL_747944|202  ···················· ···················· ···················· ···················· ···················· ····················
AA  5652 hCoV-19/Denmark/DCGC-20035/2020|EPI_ISL_747945|202  ···················· ···················· ···················· ···················· ···················· ····················
AA  5653 hCoV-19/Denmark/DCGC-20040/2020|EPI_ISL_747946|202  ···················· ···················· ···················· ···················· ···················· ····················
AA  5654 hCoV-19/Denmark/DCGC-20041/2020|EPI_ISL_747947|202  ···················· ···················· ···················· ···················· ···················· ····················
AA  5655 hCoV-19/Denmark/DCGC-20042/2020|EPI_ISL_747948|202  ···················· ···················· ···················· ···················· ···················· ····················
AA  5656 hCoV-19/Denmark/DCGC-20044/2020|EPI_ISL_747949|202  ···················· ···················· ···················· ···················· ···················· ····················
AA  5657 hCoV-19/Denmark/DCGC-20047/2020|EPI_ISL_747950|202  ···················· ···················· ···················· ···················· ···················· ····················
AA  5658 hCoV-19/Denmark/DCGC-20048/2020|EPI_ISL_747951|202  ···················· ···················· ···················· ···················· ···················· ····················
AA  5659 hCoV-19/Denmark/DCGC-20051/2020|EPI_ISL_747952|202  ···················· ···················· ···················· ···················· ···················· ····················
AA  5660 hCoV-19/Denmark/DCGC-20053/2020|EPI_ISL_747953|202  ···················· ···················· ···················· ···················· ···················· ····················
AA  5661 hCoV-19/Denmark/DCGC-20054/2020|EPI_ISL_747954|202  ···················· ···················· ···················· ···················· ···················· ····················
AA  5662 hCoV-19/Denmark/DCGC-20056/2020|EPI_ISL_747955|202  ···················· ···················· ···················· ···················· ···················· ····················
AA  5663 hCoV-19/Denmark/DCGC-20058/2020|EPI_ISL_747956|202  ···················· ···················· ···················· ···················· ···················· ····················
AA  5664 hCoV-19/Denmark/DCGC-20059/2020|EPI_ISL_747957|202  ···················· ···················· ···················· ···················· ···················· ····················
AA  5665 hCoV-19/Denmark/DCGC-20060/2020|EPI_ISL_747958|202  ···················· ···················· ···················· ···················· ···················· ····················
AA  5666 hCoV-19/Denmark/DCGC-20061/2020|EPI_ISL_747959|202  ···················· ···················· ···················· ···················· ···················· ····················
AA  5667 hCoV-19/Denmark/DCGC-20062/2020|EPI_ISL_747960|202  ···················· ···················· ···················· ···················· ···················· ····················
AA  5668 hCoV-19/Denmark/DCGC-20063/2020|EPI_ISL_747961|202  ···················· ···················· ···················· ···················· ···················· ····················
AA  5669 hCoV-19/Denmark/DCGC-20066/2020|EPI_ISL_747962|202  ···················· ···················· ···················· ···················· ···················· ····················
AA  5670 hCoV-19/Denmark/DCGC-20068/2020|EPI_ISL_747963|202  ···················· ···················· ···················· ···················· ···················· ····················
AA  5671 hCoV-19/Denmark/DCGC-20069/2020|EPI_ISL_747964|202  ···················· ···················· ···················· ···················· ···················· ····················
AA  5672 hCoV-19/Denmark/DCGC-20074/2020|EPI_ISL_747965|202  ···················· ···················· ···················· ···················· ···················· ····················
AA  5673 hCoV-19/Denmark/DCGC-20075/2020|EPI_ISL_747966|202  ···················· ···················· ···················· ···················· ···················· ····················
AA  5674 hCoV-19/Denmark/DCGC-20077/2020|EPI_ISL_747967|202  ···················· ···················· ···················· ···················· ···················· ····················
AA  5675 hCoV-19/Denmark/DCGC-20078/2020|EPI_ISL_747968|202  ···················· ···················· ···················· ···················· ···················· ····················
AA  5676 hCoV-19/Denmark/DCGC-20079/2020|EPI_ISL_747969|202  ···················· ···················· ···················· ···················· ···················· ····················
AA  5677 hCoV-19/Denmark/DCGC-20081/2020|EPI_ISL_747970|202  ···················· ···················· ···················· ···················· ···················· ····················
AA  5678 hCoV-19/Denmark/DCGC-20084/2020|EPI_ISL_747971|202  ···················· ···················· ···················· ···················· ···················· ····················
AA  5679 hCoV-19/Denmark/DCGC-20086/2020|EPI_ISL_747972|202  ···················· ···················· ···················· ···················· ···················· ····················
AA  5680 hCoV-19/Denmark/DCGC-20087/2020|EPI_ISL_747973|202  ···················· ···················· ···················· ···················· ···················· ····················
AA  5681 hCoV-19/Denmark/DCGC-20088/2020|EPI_ISL_747974|202  ···················· ···················· ···················· ···················· ···················· ····················
AA  5682 hCoV-19/Denmark/DCGC-20089/2020|EPI_ISL_747975|202  ···················· ···················· ···················· ···················· ···················· ····················
AA  5683 hCoV-19/Denmark/DCGC-20090/2020|EPI_ISL_747976|202  ···················· ···················· ···················· ···················· ···················· ····················
AA  5684 hCoV-19/Denmark/DCGC-20093/2020|EPI_ISL_747977|202  ···················· ···················· ···················· ···················· ···················· ····················
AA  5685 hCoV-19/Denmark/DCGC-20094/2020|EPI_ISL_747978|202  ···················· ···················· ···················· ···················· ···················· ····················
AA  5686 hCoV-19/Denmark/DCGC-20095/2020|EPI_ISL_747979|202  ···················· ···················· ···················· ···················· ···················· ····················
AA  5687 hCoV-19/Denmark/DCGC-20097/2020|EPI_ISL_747980|202  ···················· ···················· ···················· ···················· ···················· ····················
AA  5688 hCoV-19/Denmark/DCGC-20098/2020|EPI_ISL_747981|202  ···················· ···················· ···················· ···················· ···················· ····················
```

| | | | | | | | | |
|---|---|---|---|---|---|---|---|---|
| AA | 5689 | hCoV-19/Denmark/DCGC-20099/2020\|EPI_ISL_747982\|202 | ···················· | ···················· | ····················· | ···················· | ···················· | ···················· |
| AA | 5690 | hCoV-19/Denmark/DCGC-20102/2020\|EPI_ISL_747983\|202 | ···················· | ···················· | ····················· | ···················· | ···················· | ···················· |
| AA | 5691 | hCoV-19/Denmark/DCGC-20103/2020\|EPI_ISL_747984\|202 | ···················· | ···················· | ····················· | ···················· | ···················· | ···················· |
| AA | 5692 | hCoV-19/Denmark/DCGC-20105/2020\|EPI_ISL_747985\|202 | ···················· | ···················· | ····················· | ···················· | ···················· | ···················· |
| AA | 5693 | hCoV-19/Denmark/DCGC-20107/2020\|EPI_ISL_747986\|202 | ···················· | ···················· | ····················· | ···················· | ···················· | ···················· |
| AA | 5694 | hCoV-19/Denmark/DCGC-20108/2020\|EPI_ISL_747987\|202 | ···················· | ···················· | ····················· | ···················· | ···················· | ···················· |
| AA | 5695 | hCoV-19/Denmark/DCGC-20110/2020\|EPI_ISL_747988\|202 | ···················· | ···················· | ····················· | ···················· | ···················· | ···················· |
| AA | 5696 | hCoV-19/Denmark/DCGC-20111/2020\|EPI_ISL_747989\|202 | ···················· | ···················· | ····················· | ···················· | ···················· | ···················· |
| AA | 5697 | hCoV-19/Denmark/DCGC-20112/2020\|EPI_ISL_747990\|202 | ···················· | ·A·················· | ····················· | ···················· | ···················· | ···················· |
| AA | 5698 | hCoV-19/Denmark/DCGC-20113/2020\|EPI_ISL_747991\|202 | ···················· | ···················· | ····················· | ···················· | ···················· | ···················· |
| AA | 5699 | hCoV-19/Denmark/DCGC-20116/2020\|EPI_ISL_747992\|202 | ···················· | ···················· | ····················· | ···················· | ···················· | ···················· |
| AA | 5700 | hCoV-19/Denmark/DCGC-20117/2020\|EPI_ISL_747993\|202 | ···················· | ···················· | ····················· | ···················· | ···················· | ···················· |
| AA | 5701 | hCoV-19/Denmark/DCGC-20119/2020\|EPI_ISL_747994\|202 | ···················· | ···················· | ····················· | ···················· | ···················· | ···················· |
| AA | 5702 | hCoV-19/Denmark/DCGC-20122/2020\|EPI_ISL_747995\|202 | ···················· | ···················· | ····················· | ···················· | ···················· | ···················· |
| AA | 5703 | hCoV-19/Denmark/DCGC-20123/2020\|EPI_ISL_747996\|202 | ···················· | ···················· | ····················· | ···················· | ···················· | ···················· |
| AA | 5704 | hCoV-19/Denmark/DCGC-20124/2020\|EPI_ISL_747997\|202 | ···················· | ···················· | ····················· | ···················· | ···················· | ···················· |
| AA | 5705 | hCoV-19/Denmark/DCGC-20126/2020\|EPI_ISL_747998\|202 | ···················· | ···················· | ····················· | ···················· | ···················· | ···················· |
| AA | 5706 | hCoV-19/Denmark/DCGC-20127/2020\|EPI_ISL_747999\|202 | ···················· | ···················· | ····················· | ···················· | ···················· | ···················· |
| AA | 5707 | hCoV-19/Denmark/DCGC-17709/2020\|EPI_ISL_751159\|202 | ···················· | ···················· | ····················· | ···················· | ···················· | ···················· |
| AA | 5708 | hCoV-19/Denmark/DCGC-17710/2020\|EPI_ISL_751160\|202 | ···················· | ···················· | ····················· | ···················· | ···················· | ···················· |
| AA | 5709 | hCoV-19/Denmark/DCGC-17711/2020\|EPI_ISL_751161\|202 | ···················· | ···················· | ····················· | ···················· | ···················· | ···················· |
| AA | 5710 | hCoV-19/Denmark/DCGC-17717/2020\|EPI_ISL_751162\|202 | ···················· | ···················· | ····················· | ···················· | ···················· | ···················· |
| AA | 5711 | hCoV-19/Denmark/DCGC-17718/2020\|EPI_ISL_751163\|202 | ···················· | ···················· | ····················· | ···················· | ···················· | ···················· |
| AA | 5712 | hCoV-19/Denmark/DCGC-17719/2020\|EPI_ISL_751164\|202 | ···················· | ···················· | ····················· | ···················· | ···················· | ···················· |
| AA | 5713 | hCoV-19/Denmark/DCGC-17720/2020\|EPI_ISL_751165\|202 | ···················· | ···················· | ····················· | ···················· | ···················· | ···················· |
| AA | 5714 | hCoV-19/Denmark/DCGC-17722/2020\|EPI_ISL_751166\|202 | ···················· | ···················· | ····················· | ···················· | ···················· | ···················· |
| AA | 5715 | hCoV-19/Denmark/DCGC-17724/2020\|EPI_ISL_751167\|202 | ···················· | ···················· | ····················· | ···················· | ···················· | ···················· |
| AA | 5716 | hCoV-19/Denmark/DCGC-17725/2020\|EPI_ISL_751168\|202 | ···················· | ···················· | ····················· | ···················· | ···················· | ···················· |
| AA | 5717 | hCoV-19/Denmark/DCGC-17726/2020\|EPI_ISL_751169\|202 | ···················· | ·A·················· | ····················· | ···················· | ···················· | ···A················ |
| AA | 5718 | hCoV-19/Denmark/DCGC-17728/2020\|EPI_ISL_751170\|202 | ···················· | ···················· | ····················· | ···················· | ···················· | ···················· |
| AA | 5719 | hCoV-19/Denmark/DCGC-17730/2020\|EPI_ISL_751171\|202 | ···················· | ···················· | ····················· | ···················· | ···················· | ···················· |
| AA | 5720 | hCoV-19/Denmark/DCGC-17731/2020\|EPI_ISL_751172\|202 | ···················· | ···················· | ····················· | ···················· | ···················· | ···················· |
| AA | 5721 | hCoV-19/Denmark/DCGC-17732/2020\|EPI_ISL_751173\|202 | ···················· | ···················· | ····················· | ···················· | ···················· | ···················· |
| AA | 5722 | hCoV-19/Denmark/DCGC-17733/2020\|EPI_ISL_751174\|202 | ···················· | ···················· | ····················· | ···················· | ···················· | ···················· |
| AA | 5723 | hCoV-19/Denmark/DCGC-17734/2020\|EPI_ISL_751175\|202 | ···················· | ···················· | ····················· | ···················· | ···················· | ···················· |
| AA | 5724 | hCoV-19/Denmark/DCGC-17735/2020\|EPI_ISL_751176\|202 | ···················· | ···················· | ····················· | ···················· | ···················· | ···················· |
| AA | 5725 | hCoV-19/Denmark/DCGC-17738/2020\|EPI_ISL_751177\|202 | ···················· | ···················· | ····················· | ···················· | ···················· | ···················· |
| AA | 5726 | hCoV-19/Denmark/DCGC-17739/2020\|EPI_ISL_751178\|202 | ···················· | ···················· | ····················· | ···················· | ···················· | ···················· |
| AA | 5727 | hCoV-19/Denmark/DCGC-17740/2020\|EPI_ISL_751179\|202 | ···················· | ···················· | ····················· | ···················· | ···················· | ···················· |
| AA | 5728 | hCoV-19/Denmark/DCGC-17743/2020\|EPI_ISL_751180\|202 | ···················· | ···················· | ····················· | ···················· | ···················· | ···················· |
| AA | 5729 | hCoV-19/Denmark/DCGC-17745/2020\|EPI_ISL_751181\|202 | ···················· | ···················· | ····················· | ···················· | ······T············· | ···················· |
| AA | 5730 | hCoV-19/Denmark/DCGC-17746/2020\|EPI_ISL_751182\|202 | ···················· | ···················· | ····················· | ···················· | ···················· | ···················· |
| AA | 5731 | hCoV-19/Denmark/DCGC-17747/2020\|EPI_ISL_751183\|202 | ···················· | ···················· | ····················· | ···················· | ···················· | ···················· |
| AA | 5732 | hCoV-19/Switzerland/ZH-UZH-IMV128/2020\|EPI_ISL_751 | ···················· | ···················· | ····················· | ···················· | ···················· | ···················· |
| AA | 5733 | hCoV-19/Switzerland/ZH-UZH-IMV129/2020\|EPI_ISL_751 | ···················· | ···················· | ····················· | ···················· | ···················· | ···················· |
| AA | 5734 | hCoV-19/Switzerland/LU-UZH-IMV132/2020\|EPI_ISL_751 | ···················· | ···················· | ····················· | ···················· | ···················· | ···················· |
| AA | 5735 | hCoV-19/Switzerland/LU-UZH-IMV134/2020\|EPI_ISL_751 | ···················· | ···················· | ····················· | ···················· | ···················· | ···················· |
| AA | 5736 | hCoV-19/Switzerland/LU-UZH-IMV135/2020\|EPI_ISL_751 | ···················· | ···················· | ····················· | ···················· | ···················· | ···················· |
| AA | 5737 | hCoV-19/Switzerland/LU-UZH-IMV136/2020\|EPI_ISL_751 | ···················· | ···················· | ····················· | ···················· | ···················· | ···················· |
| AA | 5738 | hCoV-19/Switzerland/LU-UZH-IMV137/2020\|EPI_ISL_751 | ···················· | ···················· | ····················· | ···················· | ···················· | ···················· |
| AA | 5739 | hCoV-19/Finland/4LS8E7/2020\|EPI_ISL_756010\|2020-08 | ···················· | ·A·················· | ····················· | ···················· | ···················· | ···A················ |
| AA | 5740 | hCoV-19/Finland/4LS8F7/2020\|EPI_ISL_756011\|2020-08 | ···················· | ·A·················· | ····················· | ···················· | ···················· | ···A················ |
| AA | 5741 | hCoV-19/Finland/4LS8H3/2020\|EPI_ISL_756012\|2020-08 | ···················· | ···················· | ····················· | ···················· | ···················· | ···················· |
| AA | 5742 | hCoV-19/Finland/4LS9F1/2020\|EPI_ISL_756013\|2020-08 | ···················· | ·A·················· | ····················· | ···················· | ···················· | ···A················ |
| AA | 5743 | hCoV-19/Finland/7LS10C3/2020\|EPI_ISL_756014\|2020-1 | ···················· | ·A·················· | ····················· | ···················· | ···················· | ···A················ |
| AA | 5744 | hCoV-19/Finland/7LS10E9/2020\|EPI_ISL_756015\|2020-1 | ···················· | ·A·················· | ····················· | ···················· | ···················· | ···A················ |
| AA | 5745 | hCoV-19/Finland/7LS11AA7/2020\|EPI_ISL_756016\|2020- | ···················· | ·A·················· | ····················· | ···················· | ···················· | ···A················ |
| AA | 5746 | hCoV-19/Finland/7LS11AC3/2020\|EPI_ISL_756017\|2020- | ···················· | ···················· | ····················· | ···················· | ···················· | ···················· |
| AA | 5747 | hCoV-19/Finland/7LS11AE7/2020\|EPI_ISL_756018\|2020- | ···················· | ···················· | ····················· | ···················· | ···················· | ···················· |
| AA | 5748 | hCoV-19/Finland/7LS11AF1/2020\|EPI_ISL_756019\|2020- | ···················· | ·A·················· | ····················· | ···················· | ···················· | ···A················ |
| AA | 5749 | hCoV-19/Finland/7LS11AF3/2020\|EPI_ISL_756020\|2020- | ···················· | ···················· | ····················· | ···················· | ···················· | ···················· |
| AA | 5750 | hCoV-19/Finland/7LS12AB4/2020\|EPI_ISL_756021\|2020- | ···················· | ·A·················· | ····················· | ···················· | ···················· | ···A················ |
| AA | 5751 | hCoV-19/Finland/7LS13C3/2020\|EPI_ISL_756022\|2020-1 | ···················· | ···················· | ····················· | ···················· | ···················· | ···················· |
| AA | 5752 | hCoV-19/Finland/7LS13D9/2020\|EPI_ISL_756023\|2020-1 | ···················· | ···················· | ····················· | ···················· | ···················· | ···················· |
| AA | 5753 | hCoV-19/Finland/7LS13H11/2020\|EPI_ISL_756024\|2020- | ···················· | ·A·················· | ····················· | ···················· | ···················· | ···A················ |
| AA | 5754 | hCoV-19/Finland/7LS15AD9/2020\|EPI_ISL_756025\|2020- | ···················· | ·A·················· | ····················· | ···················· | ···················· | ···A················ |
| AA | 5755 | hCoV-19/Finland/7LS17D7/2020\|EPI_ISL_756026\|2020-1 | ···················· | ·A·················· | ····················· | ···················· | ···················· | ···A················ |
| AA | 5756 | hCoV-19/Finland/7LS19E9/2020\|EPI_ISL_756027\|2020-1 | ···················· | ···················· | ····················· | ···················· | ···················· | ···················· |
| AA | 5757 | hCoV-19/Finland/7LS22D11/2020\|EPI_ISL_756028\|2020- | ···················· | ···················· | ····················· | ···················· | ···················· | ···················· |
| AA | 5758 | hCoV-19/Finland/7LS23E7/2020\|EPI_ISL_756029\|2020-1 | ···················· | ·A·················· | ····················· | ···················· | ···················· | ···A················ |
| AA | 5759 | hCoV-19/Finland/7LS2AA1/2020\|EPI_ISL_756030\|2020-1 | ···················· | ···················· | ····················· | ···················· | ···················· | ···················· |
| AA | 5760 | hCoV-19/Finland/7LS2AG5/2020\|EPI_ISL_756031\|2020-1 | ···················· | ···················· | ····················· | ···················· | ···················· | ···················· |
| AA | 5761 | hCoV-19/Finland/7LS2AG8/2020\|EPI_ISL_756032\|2020-1 | ···················· | ···················· | ····················· | ···················· | ···················· | ···················· |
| AA | 5762 | hCoV-19/Finland/7LS3B3/2020\|EPI_ISL_756033\|2020-10 | ···················· | ·A·················· | ····················· | ···················· | ···················· | ···A················ |
| AA | 5763 | hCoV-19/Finland/7LS3BG5/2020\|EPI_ISL_756034\|2020-1 | ···················· | ·A·················· | ····················· | ···················· | ···················· | ···A················ |
| AA | 5764 | hCoV-19/Finland/7LS3C6/2020\|EPI_ISL_756035\|2020-10 | ···················· | ·A·················· | ····················· | ···················· | ···················· | ···A················ |
| AA | 5765 | hCoV-19/Finland/7LS4AD8/2020\|EPI_ISL_756036\|2020-1 | ···················· | ·A·················· | ····················· | ···················· | ···················· | ···A················ |
| AA | 5766 | hCoV-19/Finland/7LS4AH2/2020\|EPI_ISL_756037\|2020-1 | ···················· | ·A·················· | ····················· | ···················· | ···················· | ···A················ |
| AA | 5767 | hCoV-19/Finland/7LS4AH5/2020\|EPI_ISL_756038\|2020-1 | ···················· | ·A·················· | ····················· | ···················· | ···················· | ···A················ |
| AA | 5768 | hCoV-19/Finland/7LS5D11/2020\|EPI_ISL_756039\|2020-1 | ···················· | ···················· | ····················· | ···················· | ···················· | ···················· |

| | | | | | | | |
|---|---|---|---|---|---|---|---|
| AA | 5769 hCoV-19/Finland/7LS6AC3/2020\|EPI_ISL_756040\|2020-1 | ···················· | ·A·················· | ···················· | ···················· | ···················· | ··A················· |
| AA | 5770 hCoV-19/Finland/7LS7A11/2020\|EPI_ISL_756041\|2020-1 | ···················· | ···················· | ···················· | ···················· | ···················· | ···················· |
| AA | 5771 hCoV-19/Finland/7LS7G7/2020\|EPI_ISL_756042\|2020-10 | ···················· | ···················· | ···················· | ···················· | ···················· | ···················· |
| AA | 5772 hCoV-19/Finland/7LS8B9/2020\|EPI_ISL_756043\|2020-10 | ···················· | ···················· | ···················· | ···················· | ···················· | ···················· |
| AA | 5773 hCoV-19/Finland/7LS9A9/2020\|EPI_ISL_756044\|2020-10 | ···················· | ·A·················· | ···················· | ···················· | ···················· | ··A················· |
| AA | 5774 hCoV-19/Finland/5LS15H5/2020\|EPI_ISL_756045\|2020-1 | ···················· | ···················· | ···················· | ···················· | ···················· | ···················· |
| AA | 5775 hCoV-19/Finland/5LS16H9/2020\|EPI_ISL_756046\|2020-1 | ···················· | ···················· | ···················· | ···················· | ···················· | ···················· |
| AA | 5776 hCoV-19/Finland/5LS17A5/2020\|EPI_ISL_756047\|2020-1 | ···················· | ···················· | ···················· | ···················· | ···················· | ···················· |
| AA | 5777 hCoV-19/Finland/5LS18A5/2020\|EPI_ISL_756048\|2020-1 | ···················· | ·A·················· | ···················· | ···················· | ···················· | ··A················· |
| AA | 5778 hCoV-19/Finland/5LS18B5/2020\|EPI_ISL_756049\|2020-1 | ···················· | ···················· | ···················· | ···················· | ···················· | ···················· |
| AA | 5779 hCoV-19/Finland/17MR10cE2/2020\|EPI_ISL_757300\|2020 | ···················· | ···················· | ···················· | ···················· | ···················· | ···················· |
| AA | 5780 hCoV-19/Finland/17MR10cF11/2020\|EPI_ISL_757301\|202 | ···················· | ···················· | ···················· | ···················· | ···················· | ···················· |
| AA | 5781 hCoV-19/Finland/17MR10cH11/2020\|EPI_ISL_757302\|202 | ···················· | ···················· | ···················· | ···················· | ···················· | ···················· |
| AA | 5782 hCoV-19/Finland/17MR10cH3/2020\|EPI_ISL_757303\|2020 | ···················· | ···················· | ···················· | ···················· | ···················· | ···················· |
| AA | 5783 hCoV-19/Finland/17MR11cA1/2020\|EPI_ISL_757304\|2020 | ···················· | ···················· | ···················· | ···················· | ···················· | ···················· |
| AA | 5784 hCoV-19/Finland/17MR11cB5/2020\|EPI_ISL_757305\|2020 | ···················· | ···················· | ···················· | ···················· | ···················· | ···················· |
| AA | 5785 hCoV-19/Finland/17MR11cB6/2020\|EPI_ISL_757306\|2020 | ···················· | ···················· | ···················· | ···················· | ···················· | ···················· |
| AA | 5786 hCoV-19/Finland/17MR11cB8/2020\|EPI_ISL_757307\|2020 | ···················· | ·A·················· | ···················· | ···················· | ···················· | ··A················· |
| AA | 5787 hCoV-19/Finland/17MR11cF6/2020\|EPI_ISL_757309\|2020 | ···················· | ···················· | ···················· | ···················· | ···················· | ···················· |
| AA | 5788 hCoV-19/Finland/19MR1cF7/2020\|EPI_ISL_757340\|2020- | ···················· | ···················· | ···················· | ···················· | ···················· | ···················· |
| AA | 5789 hCoV-19/Finland/19MR1cG11/2020\|EPI_ISL_757341\|2020 | ···················· | ···················· | ···················· | ···················· | ···················· | ···················· |
| AA | 5790 hCoV-19/Finland/1MR5cA1/2020\|EPI_ISL_757342\|2020-1 | ···················· | ·A·················· | ···················· | ···················· | ···················· | ··A················· |
| AA | 5791 hCoV-19/Finland/1LS10H11/2020\|EPI_ISL_757343\|2020- | ···················· | ···················· | ···················· | ···················· | ···················· | ···················· |
| AA | 5792 hCoV-19/Finland/1LS11G3/2020\|EPI_ISL_757344\|2020-1 | ···················· | ···················· | ···················· | ···················· | ···················· | ···················· |
| AA | 5793 hCoV-19/Finland/1LS11H2/2020\|EPI_ISL_757345\|2020-1 | ···················· | ···················· | ···················· | ···················· | ···················· | ···················· |
| AA | 5794 hCoV-19/Finland/1LS11H9/2020\|EPI_ISL_757346\|2020-1 | ···················· | ·A·················· | ···················· | ···················· | ···················· | ··A················· |
| AA | 5795 hCoV-19/Finland/1LS13B5/2020\|EPI_ISL_757347\|2020-1 | ···················· | ···················· | ···················· | ···················· | ···················· | ···················· |
| AA | 5796 hCoV-19/Finland/1LS13C7/2020\|EPI_ISL_757348\|2020-1 | ···················· | ···················· | ···················· | ···················· | ···················· | ···················· |
| AA | 5797 hCoV-19/Finland/1LS13D7/2020\|EPI_ISL_757349\|2020-1 | ···················· | ·A·················· | ···················· | ···················· | ···················· | ··A················· |
| AA | 5798 hCoV-19/Finland/1LS14C9/2020\|EPI_ISL_757350\|2020-1 | ···················· | ···················· | ···················· | ···················· | ···················· | ···················· |
| AA | 5799 hCoV-19/Finland/1LS16D10/2020\|EPI_ISL_757351\|2020- | ···················· | ···················· | ···················· | ···················· | ···················· | ···················· |
| AA | 5800 hCoV-19/Finland/1LS16E3/2020\|EPI_ISL_757352\|2020-1 | ···················· | ···················· | ···················· | ···················· | ···················· | ···················· |
| AA | 5801 hCoV-19/Finland/1LS18A9/2020\|EPI_ISL_757353\|2020-1 | ···················· | ·A·················· | ···················· | ···················· | ···················· | ··A················· |
| AA | 5802 hCoV-19/Finland/1LS1D11/2020\|EPI_ISL_757354\|2020-1 | ···················· | ···················· | ···················· | ···················· | ···················· | ···················· |
| AA | 5803 hCoV-19/Finland/1LS1D7/2020\|EPI_ISL_757355\|2020-10 | ···················· | ·A·················· | ···················· | ···················· | ···················· | ··A················· |
| AA | 5804 hCoV-19/Finland/1LS20B8/2020\|EPI_ISL_757356\|2020-1 | ···················· | ·A·················· | ···················· | ···················· | ···················· | ··A················· |
| AA | 5805 hCoV-19/Finland/1LS20C2/2020\|EPI_ISL_757357\|2020-1 | ···················· | ·A·················· | ···················· | ···················· | ···················· | ··A················· |
| AA | 5806 hCoV-19/Finland/1LS23E5/2020\|EPI_ISL_757358\|2020-1 | ···················· | ···················· | ···················· | ···················· | ···················· | ···················· |
| AA | 5807 hCoV-19/Finland/1LS4C1/2020\|EPI_ISL_757359\|2020-10 | ···················· | ·A·················· | ···················· | ···················· | ···················· | ··A················· |
| AA | 5808 hCoV-19/Finland/1LS5B5/2020\|EPI_ISL_757360\|2020-10 | ···················· | ···················· | ···················· | ···················· | ···················· | ···················· |
| AA | 5809 hCoV-19/Finland/1LS5B9/2020\|EPI_ISL_757361\|2020-10 | ···················· | ···················· | ···················· | ···················· | ···················· | ···················· |
| AA | 5810 hCoV-19/Finland/1LS6C1/2020\|EPI_ISL_757362\|2020-10 | ···················· | ···················· | ···················· | ···················· | ···················· | ···················· |
| AA | 5811 hCoV-19/Finland/1LS6F1/2020\|EPI_ISL_757363\|2020-10 | ···················· | ···················· | ···················· | ···················· | ···················· | ···················· |
| AA | 5812 hCoV-19/Finland/1LS7B6/2020\|EPI_ISL_757364\|2020-10 | ···················· | ···················· | ···················· | ···················· | ···················· | ···················· |
| AA | 5813 hCoV-19/Finland/1SS14AA1/2020\|EPI_ISL_757365\|2020- | ···················· | ·A·················· | ···················· | ···················· | ···················· | ··A················· |
| AA | 5814 hCoV-19/Finland/1SS23AH1/2020\|EPI_ISL_757366\|2020- | ···················· | ···················· | ···················· | ···················· | ···················· | ···················· |
| AA | 5815 hCoV-19/Finland/1SS6AH8/2020\|EPI_ISL_757367\|2020-0 | ···················· | ·A·················· | ···················· | ···················· | ···················· | ··A················· |
| AA | 5816 hCoV-19/Finland/20MS4A4/2020\|EPI_ISL_757368\|2020-0 | ···················· | ···················· | ···················· | ···················· | ···················· | ···················· |
| AA | 5817 hCoV-19/Finland/20MS4A7/2020\|EPI_ISL_757369\|2020-0 | ···················· | ···················· | ···················· | ···················· | ···················· | ···················· |
| AA | 5818 hCoV-19/Finland/20MS4B7/2020\|EPI_ISL_757370\|2020-0 | ···················· | ···················· | ···················· | ···················· | ···················· | ···················· |
| AA | 5819 hCoV-19/Finland/20SS11F9/2020\|EPI_ISL_757371\|2020- | ···················· | ·A·················· | ···················· | ···················· | ···················· | ··A················· |
| AA | 5820 hCoV-19/Finland/20SS11G9/2020\|EPI_ISL_757372\|2020- | ···················· | ···················· | ···················· | ···················· | ···················· | ···················· |
| AA | 5821 hCoV-19/Finland/20SS11H1/2020\|EPI_ISL_757373\|2020- | ···················· | ···················· | ···················· | ···················· | ···················· | ···················· |
| AA | 5822 hCoV-19/Finland/20SS16F1/2020\|EPI_ISL_757374\|2020- | ···················· | ·A·················· | ···················· | ···················· | ···················· | ··A················· |
| AA | 5823 hCoV-19/Finland/20SS17B1/2020\|EPI_ISL_757375\|2020- | ···················· | ·A·················· | ···················· | ···················· | ···················· | ··A················· |
| AA | 5824 hCoV-19/Finland/20SS20D11/2020\|EPI_ISL_757376\|2020 | ···················· | ·A·················· | ···················· | ···················· | ···················· | ··A················· |
| AA | 5825 hCoV-19/Finland/21ES22E3/2020\|EPI_ISL_757377\|2020- | ···················· | ···················· | ···················· | ···················· | ···················· | ···················· |
| AA | 5826 hCoV-19/Finland/21KS3H2/2020\|EPI_ISL_757378\|2020-0 | ···················· | ···················· | ···················· | ···················· | ···················· | ···················· |
| AA | 5827 hCoV-19/Finland/21KS4F3/2020\|EPI_ISL_757379\|2020-0 | ···················· | ···················· | ···················· | ···················· | ···················· | ···················· |
| AA | 5828 hCoV-19/Denmark/DCGC-22193/2020\|EPI_ISL_758606\|202 | ···················· | ···················· | ···················· | ···················· | ···················· | ···················· |
| AA | 5829 hCoV-19/Denmark/DCGC-22195/2020\|EPI_ISL_758607\|202 | ···················· | ···················· | ···················· | ···················· | ···················· | ···················· |
| AA | 5830 hCoV-19/Denmark/DCGC-22198/2020\|EPI_ISL_758610\|202 | ···················· | ···················· | ···················· | ···················· | ···················· | ···················· |
| AA | 5831 hCoV-19/Denmark/DCGC-22201/2020\|EPI_ISL_758611\|202 | ···················· | ···················· | ···················· | ···················· | ···················· | ···················· |
| AA | 5832 hCoV-19/Denmark/DCGC-22205/2020\|EPI_ISL_758612\|202 | ···················· | ···················· | ···················· | ···················· | ···················· | ···················· |
| AA | 5833 hCoV-19/Denmark/DCGC-22207/2020\|EPI_ISL_758613\|202 | ···················· | ···················· | ···················· | ···················· | ···················· | ···················· |
| AA | 5834 hCoV-19/Denmark/DCGC-22208/2020\|EPI_ISL_758614\|202 | ···················· | ···················· | ···················· | ···················· | ···················· | ···················· |
| AA | 5835 hCoV-19/Denmark/DCGC-22210/2020\|EPI_ISL_758615\|202 | ···················· | ···················· | ···················· | ···················· | ···················· | ···················· |
| AA | 5836 hCoV-19/Denmark/DCGC-22211/2020\|EPI_ISL_758616\|202 | ···················· | ···················· | ···················· | ···················· | ···················· | ···················· |
| AA | 5837 hCoV-19/Denmark/DCGC-22212/2020\|EPI_ISL_758617\|202 | ···················· | ···················· | ···················· | ···················· | ···················· | ···················· |
| AA | 5838 hCoV-19/Denmark/DCGC-22213/2020\|EPI_ISL_758618\|202 | ···················· | ···················· | ···················· | ···················· | ···················· | ···················· |
| AA | 5839 hCoV-19/Denmark/DCGC-22214/2020\|EPI_ISL_758619\|202 | ···················· | ···················· | ···················· | ···················· | ···················· | ···················· |
| AA | 5840 hCoV-19/Denmark/DCGC-22216/2020\|EPI_ISL_758620\|202 | ···················· | ···················· | ···················· | ···················· | ···················· | ···················· |
| AA | 5841 hCoV-19/Denmark/DCGC-22220/2020\|EPI_ISL_758621\|202 | ···················· | ···················· | ···················· | ···················· | ···················· | ···················· |
| AA | 5842 hCoV-19/Denmark/DCGC-22221/2020\|EPI_ISL_758622\|202 | ···················· | ···················· | ···················· | ···················· | ···················· | ···················· |
| AA | 5843 hCoV-19/Denmark/DCGC-22222/2020\|EPI_ISL_758623\|202 | ···················· | ···················· | ···················· | ···················· | ···················· | ···················· |
| AA | 5844 hCoV-19/Denmark/DCGC-22223/2020\|EPI_ISL_758624\|202 | ···················· | ···················· | ···················· | ···················· | ···················· | ···················· |
| AA | 5845 hCoV-19/Denmark/DCGC-22225/2020\|EPI_ISL_758625\|202 | ···················· | ···················· | ···················· | ···················· | ···················· | ···················· |
| AA | 5846 hCoV-19/Denmark/DCGC-22227/2020\|EPI_ISL_758626\|202 | ···················· | ···················· | ···················· | ···················· | ···················· | ···················· |
| AA | 5847 hCoV-19/Denmark/DCGC-22230/2020\|EPI_ISL_758627\|202 | ···················· | ···················· | ···················· | ···················· | ···················· | ···················· |
| AA | 5848 hCoV-19/Denmark/DCGC-22233/2020\|EPI_ISL_758628\|202 | ···················· | ···················· | ···················· | ···················· | ···················· | ···················· |

| | | | | | | | |
|---|---|---|---|---|---|---|---|
| AA | 5849 hCoV-19/Denmark/DCGC-22236/2020\|EPI_ISL_758629\|202 | ···················· | ···················· | ···················· | ···················· | ···················· | ···················· |
| AA | 5850 hCoV-19/Denmark/DCGC-22239/2020\|EPI_ISL_758630\|202 | ···················· | ···················· | ···················· | ···················· | ···················· | ···················· |
| AA | 5851 hCoV-19/Denmark/DCGC-22243/2020\|EPI_ISL_758631\|202 | ···················· | ···················· | ···················· | ···················· | ···················· | ···················· |
| AA | 5852 hCoV-19/Denmark/DCGC-22244/2020\|EPI_ISL_758632\|202 | ···················· | ···················· | ···················· | ···················· | ···················· | ···················· |
| AA | 5853 hCoV-19/Denmark/DCGC-22245/2020\|EPI_ISL_758633\|202 | ···················· | ···················· | ···················· | ···················· | ···················· | ···················· |
| AA | 5854 hCoV-19/Denmark/DCGC-22246/2020\|EPI_ISL_758634\|202 | ···················· | ···················· | ···················· | ···················· | ···················· | ···················· |
| AA | 5855 hCoV-19/Denmark/DCGC-22247/2020\|EPI_ISL_758635\|202 | ···················· | ···················· | ···················· | ···················· | ···················· | ···················· |
| AA | 5856 hCoV-19/Denmark/DCGC-22252/2020\|EPI_ISL_758636\|202 | ···················· | ···················· | ···················· | ···················· | ···················· | ···················· |
| AA | 5857 hCoV-19/Denmark/DCGC-22254/2020\|EPI_ISL_758637\|202 | ···················· | ···················· | ···················· | ···················· | ···················· | ···················· |
| AA | 5858 hCoV-19/Denmark/DCGC-22255/2020\|EPI_ISL_758638\|202 | ···················· | ···················· | ···················· | ···················· | ···················· | ···················· |
| AA | 5859 hCoV-19/Denmark/DCGC-22311/2020\|EPI_ISL_758670\|202 | ···················· | ···················· | ···················· | ···················· | ···················· | ···················· |
| AA | 5860 hCoV-19/Denmark/DCGC-22312/2020\|EPI_ISL_758671\|202 | ···················· | ···················· | ···················· | ···················· | ···················· | ···················· |
| AA | 5861 hCoV-19/Denmark/DCGC-22314/2020\|EPI_ISL_758672\|202 | ···················· | ···················· | ···················· | ···················· | ···················· | ···················· |
| AA | 5862 hCoV-19/Denmark/DCGC-22315/2020\|EPI_ISL_758673\|202 | ···················· | ···················· | ···················· | ···················· | ···················· | ···················· |
| AA | 5863 hCoV-19/Denmark/DCGC-22317/2020\|EPI_ISL_758674\|202 | ···················· | ···················· | ···················· | ···················· | ···················· | ···················· |
| AA | 5864 hCoV-19/Denmark/DCGC-22318/2020\|EPI_ISL_758675\|202 | ···················· | ···················· | ···················· | ···················· | ···················· | ···················· |
| AA | 5865 hCoV-19/Denmark/DCGC-22319/2020\|EPI_ISL_758676\|202 | ···················· | ···················· | ···················· | ···················· | ···················· | ···················· |
| AA | 5866 hCoV-19/Denmark/DCGC-22323/2020\|EPI_ISL_758677\|202 | ···················· | ···················· | ···················· | ···················· | ···················· | ···················· |
| AA | 5867 hCoV-19/Denmark/DCGC-22324/2020\|EPI_ISL_758678\|202 | ···················· | ···················· | ···················· | ···················· | ···················· | ···················· |
| AA | 5868 hCoV-19/Denmark/DCGC-22325/2020\|EPI_ISL_758679\|202 | ···················· | ···················· | ···················· | ···················· | ···················· | ···················· |
| AA | 5869 hCoV-19/Denmark/DCGC-22328/2020\|EPI_ISL_758680\|202 | ···················· | ···················· | ···················· | ···················· | ···················· | ···················· |
| AA | 5870 hCoV-19/Denmark/DCGC-22330/2020\|EPI_ISL_758681\|202 | ···················· | ···················· | ···················· | ···················· | ···················· | ···················· |
| AA | 5871 hCoV-19/Denmark/DCGC-22331/2020\|EPI_ISL_758682\|202 | ···················· | ···················· | ···················· | ···················· | ···················· | ···················· |
| AA | 5872 hCoV-19/Denmark/DCGC-22332/2020\|EPI_ISL_758683\|202 | ···················· | ·A·················· | ···················· | ···················· | ···················· | ··A················· |
| AA | 5873 hCoV-19/Denmark/DCGC-22333/2020\|EPI_ISL_758684\|202 | ···················· | ···················· | ···················· | ···················· | ···················· | ···················· |
| AA | 5874 hCoV-19/Denmark/DCGC-22335/2020\|EPI_ISL_758685\|202 | ···················· | ···················· | ···················· | ···················· | ···················· | ···················· |
| AA | 5875 hCoV-19/Denmark/DCGC-22336/2020\|EPI_ISL_758686\|202 | ···················· | ···················· | ···················· | ···················· | ···················· | ···················· |
| AA | 5876 hCoV-19/Denmark/DCGC-22337/2020\|EPI_ISL_758687\|202 | ···················· | ···················· | ···················· | ···················· | ···················· | ···················· |
| AA | 5877 hCoV-19/Denmark/DCGC-22340/2020\|EPI_ISL_758688\|202 | ···················· | ···················· | ···················· | ···················· | ···················· | ···················· |
| AA | 5878 hCoV-19/Denmark/DCGC-22341/2020\|EPI_ISL_758689\|202 | ···················· | ···················· | ···················· | ···················· | ···················· | ···················· |
| AA | 5879 hCoV-19/Denmark/DCGC-22342/2020\|EPI_ISL_758690\|202 | ···················· | ···················· | ···················· | ···················· | ···················· | ···················· |
| AA | 5880 hCoV-19/Denmark/DCGC-22343/2020\|EPI_ISL_758691\|202 | ···················· | ·A·················· | ···················· | ···················· | ···················· | ··A················· |
| AA | 5881 hCoV-19/Denmark/DCGC-22344/2020\|EPI_ISL_758692\|202 | ···················· | ···················· | ···················· | ···················· | ···················· | ···················· |
| AA | 5882 hCoV-19/Denmark/DCGC-22345/2020\|EPI_ISL_758693\|202 | ···················· | ···················· | ···················· | ···················· | ···················· | ···················· |
| AA | 5883 hCoV-19/Denmark/DCGC-22346/2020\|EPI_ISL_758694\|202 | ···················· | ···················· | ···················· | ···················· | ···················· | ···················· |
| AA | 5884 hCoV-19/Denmark/DCGC-22347/2020\|EPI_ISL_758695\|202 | ···················· | ···················· | ···················· | ···················· | ···················· | ···················· |
| AA | 5885 hCoV-19/Denmark/DCGC-22350/2020\|EPI_ISL_758696\|202 | ···················· | ···················· | ···················· | ···················· | ···················· | ···················· |
| AA | 5886 hCoV-19/Denmark/DCGC-22351/2020\|EPI_ISL_758697\|202 | ···················· | ·A·················· | ···················· | ···················· | ···················· | ··A················· |
| AA | 5887 hCoV-19/Denmark/DCGC-22352/2020\|EPI_ISL_758698\|202 | ···················· | ···················· | ···················· | ···················· | ···················· | ···················· |
| AA | 5888 hCoV-19/Denmark/DCGC-22353/2020\|EPI_ISL_758699\|202 | ···················· | ···················· | ···················· | ···················· | ···················· | ···················· |
| AA | 5889 hCoV-19/Switzerland/LU-UZH-IMV138/2020\|EPI_ISL_751 | ···················· | ···················· | ···················· | ···················· | ···················· | ···················· |
| AA | 5890 hCoV-19/Saudi Arabia/KAUST-MADINAH1653/2020\|EPI_IS | ···················· | ···················· | ···················· | ···················· | ···················· | ···················· |
| AA | 5891 hCoV-19/Saudi Arabia/KAUST-MADINAH22/2020\|EPI_ISL_ | ···················· | ···················· | ···················· | ···················· | ···················· | ···················· |
| AA | 5892 hCoV-19/Saudi Arabia/KAUST-MAKKAH1505/2020\|EPI_ISL | ···················· | ···················· | ···················· | ···················· | ···················· | ···················· |
| AA | 5893 hCoV-19/India/DL-IGIB11304887/2020\|EPI_ISL_751245\| | ···················· | ·A·················· | ···················· | ···················· | ···················· | ··A················· |
| AA | 5894 hCoV-19/India/DL-IGIB11305269/2020\|EPI_ISL_751246\| | ···················· | ···················· | ···················· | ···················· | ···················· | ···················· |
| AA | 5895 hCoV-19/India/DL-IGIB11303565/2020\|EPI_ISL_751277\| | ···················· | ·A·················· | ···················· | ···················· | ···················· | ··A················· |
| AA | 5896 hCoV-19/India/DL-IGIB11304410/2020\|EPI_ISL_751278\| | ···················· | ·A·················· | ···················· | ···················· | ···················· | ··A················· |
| AA | 5897 hCoV-19/India/DL-IGIB11304579/2020\|EPI_ISL_751279\| | ···················· | ···················· | ···················· | ···················· | ···················· | ···················· |
| AA | 5898 hCoV-19/Ireland/KE-NVRL-77IRL65598/2020\|EPI_ISL_75 | ···················· | ···················· | ···················· | ···················· | ···················· | ···················· |
| AA | 5899 hCoV-19/Ireland/KE-NVRL-77IRL65631/2020\|EPI_ISL_75 | ···················· | ···················· | ···················· | ···················· | ···················· | ···················· |
| AA | 5900 hCoV-19/Ireland/D-NVRL-77IRL69349/2020\|EPI_ISL_752 | ···················· | ···················· | ···················· | ···················· | ···················· | ···················· |
| AA | 5901 hCoV-19/Ireland/D-NVRL-77IRL69365/2020\|EPI_ISL_752 | ···················· | ···················· | ···················· | ···················· | ···················· | ···················· |
| AA | 5902 hCoV-19/Ireland/D-NVRL-77IRL69369/2020\|EPI_ISL_752 | ···················· | ···················· | ···················· | ···················· | ···················· | ···················· |
| AA | 5903 hCoV-19/Ireland/MH-NVRL-77IRL69686/2020\|EPI_ISL_75 | ···················· | ···················· | ···················· | ···················· | ···················· | ···················· |
| AA | 5904 hCoV-19/Ireland/WW-NVRL-77IRL69699/2020\|EPI_ISL_75 | ···················· | ···················· | ···················· | ···················· | ···················· | ···················· |
| AA | 5905 hCoV-19/Ireland/LD-NVRL-77IRL69816/2020\|EPI_ISL_75 | ···················· | ···················· | ···················· | ···················· | ···················· | ···················· |
| AA | 5906 hCoV-19/Ireland/MH-NVRL-77IRL69897/2020\|EPI_ISL_75 | ···················· | ···················· | ···················· | ···················· | ···················· | ···················· |
| AA | 5907 hCoV-19/Ireland/MH-NVRL-77IRL71270/2020\|EPI_ISL_75 | ···················· | ···················· | ···················· | ···················· | ···················· | ···················· |
| AA | 5908 hCoV-19/Ireland/MN-NVRL-77IRL72276/2020\|EPI_ISL_75 | ···················· | ···················· | ···················· | ···················· | ···················· | ···················· |
| AA | 5909 hCoV-19/Ireland/MH-NVRL-77IRL72666/2020\|EPI_ISL_75 | ···················· | ···················· | ···················· | ···················· | ···················· | ···················· |
| AA | 5910 hCoV-19/Ireland/MH-NVRL-77IRL72674/2020\|EPI_ISL_75 | ···················· | ···················· | ···················· | ···················· | ···················· | ···················· |
| AA | 5911 hCoV-19/Ireland/MH-NVRL-77IRL72701/2020\|EPI_ISL_75 | ···················· | ···················· | ···················· | ···················· | ···················· | ···················· |
| AA | 5912 hCoV-19/Ireland/MH-NVRL-77IRL72842/2020\|EPI_ISL_75 | ···················· | ···················· | ···················· | ···················· | ···················· | ···················· |
| AA | 5913 hCoV-19/Ireland/MN-NVRL-77IRL73367/2020\|EPI_ISL_75 | ···················· | ···················· | ···················· | ···················· | ···················· | ···················· |
| AA | 5914 hCoV-19/Ireland/LH-NVRL-77IRL73961/2020\|EPI_ISL_75 | ···················· | ···················· | ···················· | ···················· | ···················· | ···················· |
| AA | 5915 hCoV-19/Ireland/CO-NVRL-77IRL74089/2020\|EPI_ISL_75 | ···················· | ···················· | ···················· | ···················· | ···················· | ···················· |
| AA | 5916 hCoV-19/Ireland/WD-NVRL-77IRL74386/2020\|EPI_ISL_75 | ···················· | ···················· | ···················· | ···················· | ···················· | ···················· |
| AA | 5917 hCoV-19/Ireland/CO-NVRL-77IRL76087/2020\|EPI_ISL_75 | ···················· | ···················· | ···················· | ···················· | ···················· | ···················· |
| AA | 5918 hCoV-19/Ireland/KK-NVRL-78IRL61331/2020\|EPI_ISL_75 | ···················· | ···················· | ···················· | ···················· | ···················· | ···················· |
| AA | 5919 hCoV-19/Ireland/TA-NVRL-78IRL62510/2020\|EPI_ISL_75 | ···················· | ···················· | ···················· | ···················· | ···················· | ···················· |
| AA | 5920 hCoV-19/Ireland/LH-NVRL-78IRL62518/2020\|EPI_ISL_75 | ···················· | ···················· | ···················· | ···················· | ···················· | ···················· |
| AA | 5921 hCoV-19/Ireland/CE-NVRL-78IRL67953/2020\|EPI_ISL_75 | ···················· | ···················· | ···················· | ···················· | ···················· | ···················· |
| AA | 5922 hCoV-19/Ireland/LK-NVRL-78IRL68004/2020\|EPI_ISL_75 | ···················· | ···················· | ···················· | ···················· | ···················· | ···················· |
| AA | 5923 hCoV-19/Ireland/TA-NVRL-78IRL68393/2020\|EPI_ISL_75 | ···················· | ···················· | ···················· | ···················· | ···················· | ···················· |
| AA | 5924 hCoV-19/Ireland/MO-NVRL-78IRL68999/2020\|EPI_ISL_75 | ···················· | ···················· | ···················· | ···················· | ···················· | ···················· |
| AA | 5925 hCoV-19/Ireland/WW-NVRL-78IRL69294/2020\|EPI_ISL_75 | ···················· | ···················· | ···················· | ···················· | ···················· | ···················· |
| AA | 5926 hCoV-19/Ireland/LK-NVRL-78IRL69435/2020\|EPI_ISL_75 | ···················· | ···················· | ···················· | ···················· | ···················· | ···················· |
| AA | 5927 hCoV-19/Ireland/WW-NVRL-78IRL70744/2020\|EPI_ISL_75 | ···················· | ·A·················· | ···················· | ···················· | ···················· | ···················· |
| AA | 5928 hCoV-19/Ireland/LK-NVRL-78IRL70746/2020\|EPI_ISL_75 | ···················· | ···················· | ···················· | ···················· | ···················· | ···················· |

| | | | | | | | |
|---|---|---|---|---|---|---|---|
| AA | 5929 hCoV-19/Ireland/D-NVRL-78IRL71541/2020\|EPI_ISL_752 | ···················· | ···················· | ···················· | ···················· | ···················· | ···················· |
| AA | 5930 hCoV-19/Ireland/WX-NVRL-78IRL71542/2020\|EPI_ISL_75 | ···················· | ···················· | ···················· | ···················· | ···················· | ···················· |
| AA | 5931 hCoV-19/Ireland/LS-NVRL-78IRL80633/2020\|EPI_ISL_75 | ···················· | ···················· | ···················· | ···················· | ···················· | ···················· |
| AA | 5932 hCoV-19/Australia/SAP597/2020\|EPI_ISL_752599\|2020- | ···················· | ···················· | ···················· | ···················· | ···················· | ···················· |
| AA | 5933 hCoV-19/Germany/DE-BE-ChVir-W1301-8113/2020\|EPI_IS | ···················· | ···················· | ···················· | ···················· | ···················· | ···················· |
| AA | 5934 hCoV-19/Germany/DE-HB-ChVir-D1974-8282/2020\|EPI_IS | ···················· | ···················· | ···················· | ···················· | ···················· | ···················· |
| AA | 5935 hCoV-19/Germany/DE-BE-ChVir-D1949-8257/2020\|EPI_IS | ···················· | ···················· | ···················· | ···················· | ···················· | ···················· |
| AA | 5936 hCoV-19/Germany/DE-BE-ChVir-D1950-4422/2020\|EPI_IS | ···················· | ···················· | ···················· | ···················· | ···················· | ···················· |
| AA | 5937 hCoV-19/Germany/DE-BE-ChVir-D2013-8255/2020\|EPI_IS | ···················· | ···················· | ···················· | ···················· | ···················· | ···················· |
| AA | 5938 hCoV-19/Germany/DE-BE-ChVir-W1302-8173/2020\|EPI_IS | ···················· | ···A················ | ···················· | ···················· | ···················· | ···················· |
| AA | 5939 hCoV-19/Germany/DE-BE-ChVir-W1304-8152/2020\|EPI_IS | ···················· | ···A················ | ···················· | ···················· | ···················· | ···················· |
| AA | 5940 hCoV-19/Germany/DE-BE-ChVir-W1428-8266/2020\|EPI_IS | ···················· | ···A················ | ···················· | ···················· | ···················· | ···················· |
| AA | 5941 hCoV-19/Germany/DE-BE-ChVir-W1447-8299/2020\|EPI_IS | ···················· | ···A················ | ···················· | ···················· | ···················· | ···················· |
| AA | 5942 hCoV-19/Germany/DE-BE-ChVir-W1648-8286/2020\|EPI_IS | ···················· | ···A················ | ···················· | ···················· | ···················· | ···················· |
| AA | 5943 hCoV-19/Germany/DE-BE-ChVir-W1077-8177/2020\|EPI_IS | ···················· | ···················· | ···················· | ···················· | ···················· | ···················· |
| AA | 5944 hCoV-19/Germany/un-ChVir-W1322-5619/2020\|EPI_ISL_7 | ···················· | ···················· | ···················· | ···················· | ···················· | ···················· |
| AA | 5945 hCoV-19/Germany/DE-BB-ChVir-D934-5623/2020\|EPI_ISL | ···················· | ···················· | ···················· | ···················· | ···················· | NN·················· |
| AA | 5946 hCoV-19/Germany/DE-BB-ChVir-D935-7207/2020\|EPI_ISL | ···················· | ···················· | ···················· | ···················· | ···················· | ···················· |
| AA | 5948 hCoV-19/Germany/DE-BE-ChVir-D1977-5942/2020\|EPI_IS | ···················· | ···················· | ···················· | ···················· | ···················· | ···················· |
| AA | 5949 hCoV-19/Germany/DE-BE-ChVir-D1978-4478/2020\|EPI_IS | ···················· | ···················· | ···················· | ···················· | ···················· | ···················· |
| AA | 5950 hCoV-19/Germany/DE-BE-ChVir-D1979-4481/2020\|EPI_IS | ···················· | ···················· | ···················· | ···················· | ···················· | ···················· |
| AA | 5951 hCoV-19/Germany/DE-BE-ChVir-D1983-4479/2020\|EPI_IS | ···················· | ···················· | ···················· | ···················· | ···················· | ···················· |
| AA | 5952 hCoV-19/Germany/DE-BE-ChVir-D1984-4423/2020\|EPI_IS | ···················· | ···················· | ···················· | ···················· | ···················· | ···················· |
| AA | 5953 hCoV-19/Germany/DE-BE-ChVir-D1988-4466/2020\|EPI_IS | ···················· | ···················· | ···················· | ···················· | ···················· | ···················· |
| AA | 5954 hCoV-19/Germany/DE-BE-ChVir-D716-2070/2020\|EPI_ISL | ···················· | ···················· | ···················· | ···················· | ···················· | ···················· |
| AA | 5955 hCoV-19/Germany/DE-BE-ChVir-W1319-2159/2020\|EPI_IS | ···················· | ···················· | ···················· | ···················· | ···················· | ···················· |
| AA | 5956 hCoV-19/Germany/DE-BE-ChVir-W1323-5723/2020\|EPI_IS | ···················· | ···················· | ···················· | ···················· | ···················· | ···················· |
| AA | 5957 hCoV-19/Germany/DE-BE-ChVir-W1389-4345/2020\|EPI_IS | ···················· | ···················· | ···················· | ···················· | ···················· | ···················· |
| AA | 5958 hCoV-19/Germany/DE-BE-ChVir-W1398-4386/2020\|EPI_IS | ···················· | ···················· | ···················· | ···················· | ···················· | ···················· |
| AA | 5959 hCoV-19/Germany/DE-BE-ChVir-W2201-4473/2020\|EPI_IS | ···················· | ·A···A·············· | ···················· | ···················· | ···················· | ···················· |
| AA | 5961 hCoV-19/Germany/DE-BE-ChVir-W2207-8292/2020\|EPI_IS | ···················· | ·A···A·············· | ···················· | ···················· | ···················· | ···················· |
| AA | 5962 hCoV-19/Germany/DE-BE-ChVir-W2209-4442/2020\|EPI_IS | ···················· | ·A···A·············· | ···················· | ···················· | ···················· | ···················· |
| AA | 5963 hCoV-19/Germany/DE-BE-ChVir-W2215-4454/2020\|EPI_IS | ···················· | ·A···A·············· | ···················· | ···················· | ···················· | ···················· |
| AA | 5964 hCoV-19/Germany/DE-BE-ChVir-W2218-8296/2020\|EPI_IS | ···················· | ·A···A·············· | ···················· | ···················· | ···················· | ···················· |
| AA | 5965 hCoV-19/Germany/DE-BE-ChVir-W2219-4467/2020\|EPI_IS | ···················· | ·A···A·············· | ···················· | ···················· | ···················· | ···················· |
| AA | 5966 hCoV-19/Germany/DE-HE-ChVir-W1320-2084/2020\|EPI_IS | ···················· | ···················· | ···················· | ···················· | ···················· | ···················· |
| AA | 5967 hCoV-19/Germany/DE-HE-ChVir-W1553-8207/2020\|EPI_IS | ···················· | ···················· | ···················· | ···················· | ···················· | ···················· |
| AA | 5968 hCoV-19/Germany/DE-SN-ChVir-W1312-4386/2020\|EPI_IS | ···················· | ···················· | ···················· | ···················· | ···················· | ···················· |
| AA | 5969 hCoV-19/Germany/un-ChVir-D1912-5994/2020\|EPI_ISL_7 | ···················· | ···················· | ···················· | ···················· | ···················· | ···················· |
| AA | 5970 hCoV-19/Germany/un-ChVir-D669-5602/2020\|EPI_ISL_75 | ···················· | ···················· | ···················· | ···················· | ···················· | ···················· |
| AA | 5971 hCoV-19/Germany/un-ChVir-W1069-5703/2020\|EPI_ISL_7 | ···················· | ···················· | ···················· | ···················· | ···················· | ···················· |
| AA | 5972 hCoV-19/Germany/un-ChVir-W1071-7216/2020\|EPI_ISL_7 | ···················· | ···················· | ···················· | ···················· | ···················· | ···················· |
| AA | 5973 hCoV-19/Germany/un-ChVir-W1296-5676/2020\|EPI_ISL_7 | ···················· | ···················· | ···················· | ···················· | ···················· | ···················· |
| AA | 5974 hCoV-19/Germany/un-ChVir-W1723-4346/2020\|EPI_ISL_7 | ···················· | ···················· | ···················· | ···················· | ···················· | ···················· |
| AA | 5975 hCoV-19/Germany/DE-BB-ChVir-W1041-5644/2020\|EPI_IS | ···················· | ···················· | ···················· | ···················· | ···················· | ···················· |
| AA | 5976 hCoV-19/Germany/DE-BB-ChVir-W1052-7250/2020\|EPI_IS | ···················· | ···················· | ···················· | ···················· | ···················· | ···················· |
| AA | 5977 hCoV-19/Germany/DE-BB-ChVir-W1061-7256/2020\|EPI_IS | ···················· | ···················· | ···················· | ···················· | ···················· | ···················· |
| AA | 5978 hCoV-19/Germany/DE-BB-ChVir-W1062-7296/2020\|EPI_IS | ···················· | ···················· | ···················· | ···················· | ···················· | ···················· |
| AA | 5979 hCoV-19/Germany/DE-BB-ChVir-W1066-7201/2020\|EPI_IS | ···················· | ···················· | ···················· | ···················· | ···················· | ···················· |
| AA | 5980 hCoV-19/Germany/DE-BB-ChVir-W1067-7224/2020\|EPI_IS | ···················· | ···················· | ···················· | ···················· | ···················· | ···················· |
| AA | 5981 hCoV-19/Germany/DE-BB-ChVir-W1073-8108/2020\|EPI_IS | ···················· | ···················· | ···················· | ···················· | ···················· | ···················· |
| AA | 5982 hCoV-19/Germany/DE-BB-ChVir-W1129-7275/2020\|EPI_IS | ···················· | ···················· | ···················· | ···················· | ···················· | ···················· |
| AA | 5983 hCoV-19/Germany/DE-BB-ChVir-W1135-7242/2020\|EPI_IS | ···················· | ···················· | ···················· | ···················· | ···················· | ···················· |
| AA | 5984 hCoV-19/Germany/DE-BB-ChVir-W1144-7244/2020\|EPI_IS | ···················· | ···················· | ···················· | ···················· | ···················· | ···················· |
| AA | 5985 hCoV-19/Germany/DE-BB-ChVir-W1413-8296/2020\|EPI_IS | ···················· | ···················· | ···················· | ···················· | ···················· | ···················· |
| AA | 5986 hCoV-19/Germany/DE-BB-ChVir-W1419-8228/2020\|EPI_IS | ···················· | ···················· | ···················· | ···················· | ···················· | ···················· |
| AA | 5987 hCoV-19/Germany/DE-BB-ChVir-W1423-8226/2020\|EPI_IS | ···················· | ···················· | ···················· | ···················· | ···················· | ···················· |
| AA | 5988 hCoV-19/Germany/DE-BB-ChVir-W1426-8229/2020\|EPI_IS | ···················· | ···················· | ···················· | ···················· | ···················· | ···················· |
| AA | 5989 hCoV-19/Germany/DE-BB-ChVir-W1442-8234/2020\|EPI_IS | ···················· | ···················· | ···················· | ···················· | ···················· | ···················· |
| AA | 5990 hCoV-19/Germany/DE-BB-ChVir-W1646-6734/2020\|EPI_IS | ···················· | ···················· | ···················· | ···················· | ···················· | ···················· |
| AA | 5991 hCoV-19/Germany/DE-BB-ChVir-W2128-8294/2020\|EPI_IS | ···················· | ·A·················· | ···················· | ···················· | ···················· | ···················· |
| AA | 5992 hCoV-19/Germany/DE-BE-ChVir-D1911-8207/2020\|EPI_IS | ···················· | ···················· | ···················· | ···················· | ···················· | ···················· |
| AA | 5993 hCoV-19/Germany/DE-BE-ChVir-D1913-7445/2020\|EPI_IS | ···················· | ···················· | ···················· | ···················· | ···················· | ···················· |
| AA | 5994 hCoV-19/Germany/DE-BE-ChVir-D1914-8251/2020\|EPI_IS | ···················· | ···················· | ···················· | ···················· | ···················· | ···················· |
| AA | 5995 hCoV-19/Germany/DE-BE-ChVir-D1916-8268/2020\|EPI_IS | ···················· | ···················· | ···················· | ···················· | ·····T·············· | ···················· |
| AA | 5996 hCoV-19/Germany/DE-BE-ChVir-D1918-8214/2020\|EPI_IS | ···················· | ···················· | ···················· | ···················· | ···················· | ···················· |
| AA | 5997 hCoV-19/Germany/DE-BE-ChVir-D1919-4432/2020\|EPI_IS | ···················· | ···················· | ···················· | ···················· | ···················· | ···················· |
| AA | 5998 hCoV-19/Germany/DE-BE-ChVir-D1920-8202/2020\|EPI_IS | ···················· | ···················· | ···················· | ···················· | ···················· | ···················· |
| AA | 5999 hCoV-19/Germany/DE-BE-ChVir-D1922-4497/2020\|EPI_IS | ···················· | ···················· | ···················· | ···················· | ···················· | ···················· |
| AA | 6000 hCoV-19/Germany/DE-BE-ChVir-D1923-8213/2020\|EPI_IS | ···················· | ···················· | ···················· | ···················· | ···················· | ···················· |
| AA | 6001 hCoV-19/Germany/DE-BE-ChVir-D1924-8211/2020\|EPI_IS | ···················· | ···················· | ···················· | ···················· | ···················· | ···················· |
| AA | 6002 hCoV-19/Germany/DE-BE-ChVir-D1926-5089/2020\|EPI_IS | ···················· | ···················· | ···················· | ···················· | ···················· | ···················· |
| AA | 6003 hCoV-19/Germany/DE-BE-ChVir-D1927-7462/2020\|EPI_IS | ···················· | ···················· | ···················· | ···················· | ···················· | ···················· |
| AA | 6004 hCoV-19/Germany/DE-BE-ChVir-D1928-7459/2020\|EPI_IS | ···················· | ···················· | ···················· | ···················· | ···················· | ···················· |
| AA | 6005 hCoV-19/Germany/DE-BE-ChVir-D1930-8296/2020\|EPI_IS | ···················· | ···A················ | ···················· | ···················· | ···················· | ···················· |
| AA | 6006 hCoV-19/Germany/DE-BE-ChVir-D1932-8298/2020\|EPI_IS | ···················· | ···················· | ···················· | ···················· | ···················· | ···················· |
| AA | 6007 hCoV-19/Germany/DE-BE-ChVir-D1935-5099/2020\|EPI_IS | ···················· | ···················· | ···················· | ···················· | ···················· | ···················· |
| AA | 6008 hCoV-19/Germany/DE-BE-ChVir-D2010-8268/2020\|EPI_IS | ···················· | ···················· | ···················· | ···················· | ···················· | ···················· |
| AA | 6009 hCoV-19/Germany/DE-BE-ChVir-D2011-5016/2020\|EPI_IS | ···················· | ···················· | ···················· | ···················· | ·····T·············· | ···················· |
| AA | 6010 hCoV-19/Finland/4LS20E5/2020\|EPI_ISL_756001\|2020-0 | ···················· | ·A·················· | ···················· | ···················· | ···················· | ···A················ |

| | | | | | | | | |
|---|---|---|---|---|---|---|---|---|
| AA | 6011 | hCoV-19/Finland/4LS20F5/2020\|EPI_ISL_756002\|2020-0 | .................... | .A.................. | .................... | .................... | .................... | ...A................ |
| AA | 6012 | hCoV-19/Finland/4LS21F5/2020\|EPI_ISL_756003\|2020-0 | .................... | .................... | .................... | .................... | .................... | .................... |
| AA | 6013 | hCoV-19/Finland/4LS5A12/2020\|EPI_ISL_756004\|2020-0 | .................... | .................... | .................... | .................... | .................... | .................... |
| AA | 6014 | hCoV-19/Finland/4LS5D11/2020\|EPI_ISL_756005\|2020-0 | .................... | .................... | .................... | .................... | .................... | .................... |
| AA | 6015 | hCoV-19/Finland/4LS8A9/2020\|EPI_ISL_756007\|2020-08 | .................... | .A.................. | .................... | .................... | .................... | ...A................ |
| AA | 6016 | hCoV-19/Finland/4LS8D7/2020\|EPI_ISL_756008\|2020-08 | .................... | .A.................. | .................... | .................... | .................... | ...A................ |
| AA | 6017 | hCoV-19/Finland/4LS8E1/2020\|EPI_ISL_756009\|2020-08 | .................... | .A.................. | .................... | .................... | .................... | ...A................ |
| AA | 6018 | hCoV-19/Finland/5LS18C7/2020\|EPI_ISL_756050\|2020-1 | .................... | .A.................. | .................... | .................... | .................... | ...A................ |
| AA | 6019 | hCoV-19/Finland/5LS20C12/2020\|EPI_ISL_756051\|2020- | .................... | .A.................. | .................... | .................... | .................... | ...A................ |
| AA | 6020 | hCoV-19/Finland/5LS20C8/2020\|EPI_ISL_756052\|2020-1 | .................... | .A.................. | .................... | .................... | .................... | ...A................ |
| AA | 6021 | hCoV-19/Finland/5LS20D9/2020\|EPI_ISL_756053\|2020-1 | .................... | .A.................. | .................... | .................... | .................... | ...A................ |
| AA | 6022 | hCoV-19/Finland/5LS20H9/2020\|EPI_ISL_756054\|2020-1 | .................... | .................... | .................... | .................... | .................... | .................... |
| AA | 6023 | hCoV-19/Finland/5LS21D9/2020\|EPI_ISL_756055\|2020-1 | .................... | .................... | .................... | .................... | .................... | .................... |
| AA | 6024 | hCoV-19/Finland/5LS21E9/2020\|EPI_ISL_756056\|2020-1 | .................... | .A.................. | .................... | .................... | .................... | ...A................ |
| AA | 6025 | hCoV-19/Finland/5LS4B6/2020\|EPI_ISL_756057\|2020-10 | .................... | .................... | .................... | .................... | .................... | .................... |
| AA | 6026 | hCoV-19/Finland/5LS4D11/2020\|EPI_ISL_756058\|2020-1 | .................... | .A.................. | .................... | .................... | .................... | ...A................ |
| AA | 6027 | hCoV-19/Finland/5LS4D2/2020\|EPI_ISL_756059\|2020-10 | .................... | .A.................. | .................... | .................... | .................... | ...A................ |
| AA | 6028 | hCoV-19/Finland/5LS5A7/2020\|EPI_ISL_756060\|2020-10 | .................... | .A.................. | .................... | .................... | .................... | ...A................ |
| AA | 6029 | hCoV-19/Finland/5LS5C11/2020\|EPI_ISL_756061\|2020-1 | .................... | .................... | .................... | .................... | .................... | .................... |
| AA | 6030 | hCoV-19/Finland/5LS5E7/2020\|EPI_ISL_756062\|2020-10 | .................... | .................... | .................... | .................... | .................... | .................... |
| AA | 6031 | hCoV-19/Finland/5LS5F5/2020\|EPI_ISL_756063\|2020-10 | .................... | .................... | .................... | .................... | .................... | .................... |
| AA | 6032 | hCoV-19/Finland/5LS5G3/2020\|EPI_ISL_756064\|2020-10 | .................... | .................... | .................... | .................... | .................... | .................... |
| AA | 6033 | hCoV-19/Finland/5LS6B3/2020\|EPI_ISL_756065\|2020-10 | .................... | .A.................. | .................... | .................... | .................... | ...A................ |
| AA | 6034 | hCoV-19/Finland/5LS6H7/2020\|EPI_ISL_756066\|2020-10 | .................... | .A.................. | .................... | .................... | .................... | ...A................ |
| AA | 6035 | hCoV-19/Finland/5LS7C1/2020\|EPI_ISL_756067\|2020-10 | .................... | .A.................. | .................... | .................... | .................... | ...A................ |
| AA | 6036 | hCoV-19/Finland/5LS7G3/2020\|EPI_ISL_756068\|2020-10 | .................... | .A.................. | .................... | .................... | .................... | ...A................ |
| AA | 6037 | hCoV-19/Finland/5LS8B3/2020\|EPI_ISL_756069\|2020-10 | .................... | .................... | .................... | .................... | .................... | .................... |
| AA | 6038 | hCoV-19/Finland/5LS8D2/2020\|EPI_ISL_756070\|2020-10 | .................... | .................... | .................... | .................... | .................... | .................... |
| AA | 6039 | hCoV-19/Finland/5LS9C7/2020\|EPI_ISL_756071\|2020-10 | .................... | .................... | .................... | .................... | .................... | .................... |
| AA | 6040 | hCoV-19/Finland/6LS10B1/2020\|EPI_ISL_756072\|2020-1 | .................... | .A.................. | .................... | .................... | .................... | ...A................ |
| AA | 6041 | hCoV-19/Finland/6LS11F1/2020\|EPI_ISL_756073\|2020-1 | .................... | .................... | .................... | .................... | .................... | .................... |
| AA | 6042 | hCoV-19/Finland/6LS12E9/2020\|EPI_ISL_756074\|2020-1 | .................... | .A.................. | .................... | .................... | .................... | ...A................ |
| AA | 6043 | hCoV-19/Finland/6LS13A10/2020\|EPI_ISL_756075\|2020- | .................... | .A.................. | .................... | .................... | .................... | ...A................ |
| AA | 6044 | hCoV-19/Finland/6LS13B1/2020\|EPI_ISL_756076\|2020-1 | .................... | .................... | .................... | .................... | .................... | .................... |
| AA | 6045 | hCoV-19/Finland/6LS17C1/2020\|EPI_ISL_756077\|2020-1 | .................... | .A.................. | .................... | .................... | .................... | ...A................ |
| AA | 6046 | hCoV-19/Finland/6LS18C7/2020\|EPI_ISL_756078\|2020-1 | .................... | .................... | .................... | .................... | .................... | .................... |
| AA | 6047 | hCoV-19/Finland/6LS18D8/2020\|EPI_ISL_756079\|2020-1 | .................... | .A.................. | .................... | .................... | .................... | ...A................ |
| AA | 6048 | hCoV-19/Finland/6LS18F5/2020\|EPI_ISL_756080\|2020-1 | .................... | .................... | .................... | .................... | .................... | .................... |
| AA | 6049 | hCoV-19/Finland/6LS18F7/2020\|EPI_ISL_756081\|2020-1 | .................... | .................... | .................... | .................... | .................... | .................... |
| AA | 6050 | hCoV-19/Finland/6LS1A1/2020\|EPI_ISL_756082\|2020-10 | .................... | .A.................. | .................... | .................... | .................... | ...A................ |
| AA | 6051 | hCoV-19/Finland/6LS1H7/2020\|EPI_ISL_756083\|2020-10 | .................... | .................... | .................... | .................... | .................... | .................... |
| AA | 6052 | hCoV-19/Finland/6LS3C1/2020\|EPI_ISL_756084\|2020-10 | .................... | .A.................. | .................... | .................... | .................... | ...A................ |
| AA | 6053 | hCoV-19/Finland/6LS3C9/2020\|EPI_ISL_756085\|2020-10 | .................... | .................... | .................... | .................... | .................... | .................... |
| AA | 6054 | hCoV-19/Finland/6LS3D11/2020\|EPI_ISL_756086\|2020-1 | .................... | .................... | .................... | .................... | .................... | .................... |
| AA | 6055 | hCoV-19/Finland/6LS3H9/2020\|EPI_ISL_756087\|2020-10 | .................... | .A.................. | .................... | .................... | .................... | ...A................ |
| AA | 6056 | hCoV-19/Finland/6SS7C7/2020\|EPI_ISL_756088\|2020-09 | .................... | .A.................. | .................... | .................... | .................... | ...A................ |
| AA | 6057 | hCoV-19/Finland/6SS7H1/2020\|EPI_ISL_756089\|2020-09 | .................... | .A.................. | .................... | .................... | .................... | ...A................ |
| AA | 6058 | hCoV-19/Finland/3LS10AF1/2020\|EPI_ISL_756090\|2020- | .................... | .A.................. | .................... | .................... | .................... | ...A................ |
| AA | 6059 | hCoV-19/Finland/3LS10AF3/2020\|EPI_ISL_756091\|2020- | .................... | .A.................. | .................... | .................... | .................... | ...A................ |
| AA | 6060 | hCoV-19/Finland/3LS11AF11/2020\|EPI_ISL_756092\|2020 | .................... | .................... | .................... | .................... | .................... | .................... |
| AA | 6061 | hCoV-19/Finland/3LS12BA11/2020\|EPI_ISL_756093\|2020 | .................... | .A.................. | .................... | .................... | .................... | ...A................ |
| AA | 6062 | hCoV-19/Finland/3LS12BE5/2020\|EPI_ISL_756094\|2020- | .................... | .A.................. | .................... | .................... | .................... | ...A................ |
| AA | 6063 | hCoV-19/Finland/3LS14AG3/2020\|EPI_ISL_756095\|2020- | .................... | .................... | .................... | .................... | .................... | .................... |
| AA | 6064 | hCoV-19/Finland/3LS17G9/2020\|EPI_ISL_756096\|2020-1 | .................... | .A.................. | .................... | .................... | .................... | ...A................ |
| AA | 6065 | hCoV-19/Finland/3LS18AD6/2020\|EPI_ISL_756097\|2020- | .................... | .A.................. | .................... | .................... | .................... | ...A................ |
| AA | 6066 | hCoV-19/Finland/3LS18AD8/2020\|EPI_ISL_756098\|2020- | .................... | .A.................. | .................... | .................... | .................... | ...A................ |
| AA | 6067 | hCoV-19/Finland/3LS18AE1/2020\|EPI_ISL_756099\|2020- | .................... | .................... | .................... | .................... | .................... | .................... |
| AA | 6068 | hCoV-19/Finland/17MR11cC6/2020\|EPI_ISL_757308\|2020 | .................... | .................... | .................... | .................... | .................... | .................... |
| AA | 6069 | hCoV-19/Finland/17MR11cG5/2020\|EPI_ISL_757310\|2020 | .................... | .................... | .................... | .................... | .................... | .................... |
| AA | 6070 | hCoV-19/Finland/17MR11cG6/2020\|EPI_ISL_757311\|2020 | .................... | .................... | .................... | .................... | .................... | .................... |
| AA | 6071 | hCoV-19/Finland/17MR11cG7/2020\|EPI_ISL_757312\|2020 | .................... | .................... | .................... | .................... | .................... | .................... |
| AA | 6072 | hCoV-19/Finland/17MR11cH6/2020\|EPI_ISL_757313\|2020 | .................... | .................... | .................... | .................... | .................... | .................... |
| AA | 6073 | hCoV-19/Finland/17MR12cD12/2020\|EPI_ISL_757314\|202 | .................... | .................... | .................... | .................... | .................... | .................... |
| AA | 6074 | hCoV-19/Finland/17MR12cH11/2020\|EPI_ISL_757315\|202 | .................... | .................... | .................... | .................... | .................... | .................... |
| AA | 6075 | hCoV-19/Finland/17MR4cE10/2020\|EPI_ISL_757316\|2020 | .................... | .................... | .................... | .................... | .................... | .................... |
| AA | 6076 | hCoV-19/Finland/17MR7cE12/2020\|EPI_ISL_757317\|2020 | .................... | .................... | .................... | .................... | .................... | .................... |
| AA | 6077 | hCoV-19/Finland/18MR1cB11/2020\|EPI_ISL_757318\|2020 | .................... | .................... | .................... | .................... | .................... | .................... |
| AA | 6078 | hCoV-19/Finland/18MR1cC12/2020\|EPI_ISL_757319\|2020 | .................... | .................... | .................... | .................... | .................... | .................... |
| AA | 6079 | hCoV-19/Finland/18MR1cD7/2020\|EPI_ISL_757320\|2020- | .................... | .A.................. | .................... | .................... | .................... | ...A................ |
| AA | 6080 | hCoV-19/Finland/18MR1cG2/2020\|EPI_ISL_757321\|2020- | .................... | .................... | .................... | .................... | .................... | .................... |
| AA | 6081 | hCoV-19/Finland/18MR1cG9/2020\|EPI_ISL_757322\|2020- | .................... | .................... | .................... | .................... | .................... | .................... |
| AA | 6082 | hCoV-19/Finland/18MR2cA8/2020\|EPI_ISL_757323\|2020- | .................... | .A.................. | .................... | .................... | .................... | ...A................ |
| AA | 6083 | hCoV-19/Finland/18MR2cB10/2020\|EPI_ISL_757324\|2020 | .................... | .................... | .................... | .................... | .................... | .................... |
| AA | 6084 | hCoV-19/Finland/18MR2cF11/2020\|EPI_ISL_757325\|2020 | .................... | .................... | .................... | .................... | .................... | .................... |
| AA | 6085 | hCoV-19/Finland/18MR2cF1/2020\|EPI_ISL_757326\|2020- | .................... | .A.................. | .................... | .................... | .................... | ...A................ |
| AA | 6086 | hCoV-19/Finland/18MR2cG11/2020\|EPI_ISL_757327\|2020 | .................... | .................... | .................... | .................... | .................... | .................... |
| AA | 6087 | hCoV-19/Finland/18MR2cG9/2020\|EPI_ISL_757328\|2020- | .................... | .A.................. | .................... | .................... | .................... | ...A................ |
| AA | 6088 | hCoV-19/Finland/18MR4cF8/2020\|EPI_ISL_757329\|2020- | .................... | .................... | .................... | .................... | .................... | .................... |
| AA | 6089 | hCoV-19/Finland/18MR4cG5/2020\|EPI_ISL_757330\|2020- | .................... | .................... | .................... | .................... | .................... | .................... |
| AA | 6090 | hCoV-19/Finland/18MR5cB12/2020\|EPI_ISL_757331\|2020 | .................... | .................... | .................... | .................... | .................... | .................... |

AA 6091 hCoV-19/Finland/18MR8cB7/2020|EPI_ISL_757332|2020- ···················· ···················· ···················· ···················· ···················· ····················
AA 6092 hCoV-19/Finland/18MR8cE5/2020|EPI_ISL_757333|2020- ···················· ·A·················· ···················· ···················· ···················· ··A·················
AA 6093 hCoV-19/Finland/18MR8cE9/2020|EPI_ISL_757334|2020- ···················· ···················· ···················· ···················· ···················· ····················
AA 6094 hCoV-19/Finland/18MR9cB3/2020|EPI_ISL_757335|2020- ···················· ···················· ···················· ···················· ···················· ····················
AA 6095 hCoV-19/Finland/18MR9cC4/2020|EPI_ISL_757336|2020- ···················· ···················· ···················· ···················· ···················· ····················
AA 6096 hCoV-19/Finland/18MR9cH4/2020|EPI_ISL_757337|2020- ···················· ···················· ···················· ···················· ···················· ····················
AA 6097 hCoV-19/Finland/19MR1cA2/2020|EPI_ISL_757338|2020- ···················· ···················· ···················· ···················· ···················· ····················
AA 6098 hCoV-19/Finland/19MR1cE8/2020|EPI_ISL_757339|2020- ···················· ···················· ···················· ···················· ···················· ····················
AA 6099 hCoV-19/Finland/21SS13C8/2020|EPI_ISL_757380|2020- ···················· ···················· ···················· ···················· ···················· ····················
AA 6100 hCoV-19/Finland/21SS13G9/2020|EPI_ISL_757381|2020- ···················· ·A·················· ···················· ···················· ···················· ··A·················
AA 6101 hCoV-19/Finland/21SS4D3/2020|EPI_ISL_757382|2020-0 ···················· ···················· ···················· ···················· ···················· ····················
AA 6102 hCoV-19/Finland/22ES13AD6/2020|EPI_ISL_757383|2020 ···················· ···················· ···················· ···················· ···················· ····················
AA 6103 hCoV-19/Finland/22ES3AG1/2020|EPI_ISL_757384|2020- ···················· ···················· ···················· ···················· ···················· ····················
AA 6104 hCoV-19/Finland/22LS19D5/2020|EPI_ISL_757385|2020- ···················· ·A·················· ···················· ···················· ···················· ··A·················
AA 6105 hCoV-19/Finland/22SS11B3/2020|EPI_ISL_757386|2020- ···················· ·A·················· ···················· ···················· ···················· ··A·················
AA 6106 hCoV-19/Finland/22SS11D1/2020|EPI_ISL_757387|2020- ···················· ···················· ···················· ···················· ···················· ····················
AA 6107 hCoV-19/Finland/22SS11D7/2020|EPI_ISL_757388|2020- ···················· ·A·················· ···················· ···················· ···················· ··A·················
AA 6108 hCoV-19/Finland/22SS11E11/2020|EPI_ISL_757389|2020 ···················· ···················· ···················· ···················· ···················· ····················
AA 6109 hCoV-19/Finland/22SS11E3/2020|EPI_ISL_757390|2020- ···················· ·A·················· ···················· ···················· ···················· ····················
AA 6110 hCoV-19/Finland/22SS12E11/2020|EPI_ISL_757391|2020 ···················· ·A·················· ···················· ···················· ···················· ··A·················
AA 6111 hCoV-19/Finland/22SS4H11/2020|EPI_ISL_757392|2020- ···················· ·A·················· ···················· ···················· ···················· ··A·················
AA 6112 hCoV-19/Finland/22SS6F5/2020|EPI_ISL_757393|2020-0 ···················· ···················· ···················· ···················· ···················· ····················
AA 6113 hCoV-19/Finland/22SS9C3/2020|EPI_ISL_757394|2020-0 ···················· ···················· ···················· ···················· ···················· ····················
AA 6114 hCoV-19/Denmark/DCGC-22256/2020|EPI_ISL_758639|202 ···················· ···················· ···················· ···················· ···················· ····················
AA 6115 hCoV-19/Denmark/DCGC-22257/2020|EPI_ISL_758640|202 ···················· ···················· ···················· ···················· ···················· ····················
AA 6116 hCoV-19/Denmark/DCGC-22259/2020|EPI_ISL_758641|202 ···················· ···················· ···················· ···················· ···················· ····················
AA 6117 hCoV-19/Denmark/DCGC-22260/2020|EPI_ISL_758642|202 ···················· ···················· ···················· ···················· ···················· ····················
AA 6118 hCoV-19/Denmark/DCGC-22261/2020|EPI_ISL_758643|202 ···················· ···················· ···················· ···················· ···················· ····················
AA 6119 hCoV-19/Denmark/DCGC-22262/2020|EPI_ISL_758644|202 ···················· ···················· ···················· ···················· ···················· ····················
AA 6120 hCoV-19/Denmark/DCGC-22268/2020|EPI_ISL_758645|202 ···················· ···················· ···················· ···················· ···················· ····················
AA 6121 hCoV-19/Denmark/DCGC-22270/2020|EPI_ISL_758646|202 ···················· ···················· ··A················· ···················· ···················· ····················
AA 6122 hCoV-19/Denmark/DCGC-22272/2020|EPI_ISL_758647|202 ···················· ···················· ···················· ···················· ···················· ····················
AA 6123 hCoV-19/Denmark/DCGC-22273/2020|EPI_ISL_758648|202 ···················· ···················· ···················· ···················· ···················· ····················
AA 6124 hCoV-19/Denmark/DCGC-22275/2020|EPI_ISL_758649|202 ···················· ···················· ···················· ···················· ···················· ····················
AA 6125 hCoV-19/Denmark/DCGC-22277/2020|EPI_ISL_758650|202 ···················· ···················· ···················· ···················· ···················· ····················
AA 6126 hCoV-19/Denmark/DCGC-22279/2020|EPI_ISL_758651|202 ···················· ···················· ···················· ···················· ···················· ····················
AA 6127 hCoV-19/Denmark/DCGC-22280/2020|EPI_ISL_758652|202 ···················· ···················· ···················· ···················· ···················· ····················
AA 6128 hCoV-19/Denmark/DCGC-22281/2020|EPI_ISL_758653|202 ···················· ···················· ···················· ···················· ···················· ····················
AA 6129 hCoV-19/Denmark/DCGC-22282/2020|EPI_ISL_758654|202 ···················· ···················· ···················· ···················· ···················· ····················
AA 6130 hCoV-19/Denmark/DCGC-22283/2020|EPI_ISL_758655|202 ···················· ···················· ···················· ···················· ···················· ····················
AA 6131 hCoV-19/Denmark/DCGC-22285/2020|EPI_ISL_758656|202 ···················· ···················· ···················· ···················· ···················· ····················
AA 6132 hCoV-19/Denmark/DCGC-22286/2020|EPI_ISL_758657|202 ···················· ···················· ···················· ···················· ···················· ····················
AA 6133 hCoV-19/Denmark/DCGC-22288/2020|EPI_ISL_758658|202 ···················· ···················· ···················· ···················· ···················· ····················
AA 6134 hCoV-19/Denmark/DCGC-22290/2020|EPI_ISL_758659|202 ···················· ···················· ···················· ···················· ···················· ····················
AA 6135 hCoV-19/Denmark/DCGC-22295/2020|EPI_ISL_758660|202 ···················· ···················· ···················· ···················· ···················· ····················
AA 6136 hCoV-19/Denmark/DCGC-22296/2020|EPI_ISL_758661|202 ···················· ···················· ···················· ···················· ···················· ····················
AA 6137 hCoV-19/Denmark/DCGC-22297/2020|EPI_ISL_758662|202 ···················· ···················· ···················· ···················· ···················· ····················
AA 6138 hCoV-19/Denmark/DCGC-22298/2020|EPI_ISL_758663|202 ···················· ···················· ···················· ···················· ···················· ····················
AA 6139 hCoV-19/Denmark/DCGC-22300/2020|EPI_ISL_758664|202 ···················· ···················· ···················· ···················· ···················· ····················
AA 6140 hCoV-19/Denmark/DCGC-22301/2020|EPI_ISL_758665|202 ···················· ···················· ···················· ···················· ···················· ····················
AA 6141 hCoV-19/Denmark/DCGC-22306/2020|EPI_ISL_758666|202 ···················· ···················· ···················· ···················· ···················· ····················
AA 6142 hCoV-19/Denmark/DCGC-22307/2020|EPI_ISL_758667|202 ···················· ···················· ···················· ···················· ···················· ····················
AA 6143 hCoV-19/Denmark/DCGC-22308/2020|EPI_ISL_758668|202 ···················· ···················· ···················· ···················· ···················· ····················
AA 6144 hCoV-19/Denmark/DCGC-22309/2020|EPI_ISL_758669|202 ···················· ···················· ···················· ···················· ···················· ····················
AA 6145 hCoV-19/Saudi Arabia/KAUST-MAKKAH1534/2020|EPI_ISL ···················· ···················· ···················· ···················· ···················· ····················
AA 6146 hCoV-19/Saudi Arabia/KAUST-JEDDAH489/2020|EPI_ISL_ ···················· ···················· ···················· ···················· ···················· ····················
AA 6147 hCoV-19/Saudi Arabia/KAUST-MADINAH1669/2020|EPI_IS ···················· ·A·················· ···················· ···················· ···················· ····················
AA 6148 hCoV-19/Saudi Arabia/KAUST-MADINAH1648/2020|EPI_IS ···················· ···················· ···················· ···················· ···················· ····················
AA 6149 hCoV-19/Saudi Arabia/KAUST-MADINAH1677/2020|EPI_IS ···················· ·A·················· ···················· ···················· ···················· ····················
AA 6150 hCoV-19/Saudi Arabia/KAUST-MADINAH1497/2020|EPI_IS ···················· ···················· ···················· ···················· ···················· ··A·················
AA 6151 hCoV-19/Saudi Arabia/KAUST-MADINAH1670/2020|EPI_IS ···················· ···················· ···················· ···················· ···················· ····················
AA 6152 hCoV-19/Saudi Arabia/KAUST-MADINAH1661/2020|EPI_IS ···················· ·A·················· ···················· ···················· ···················· ··A·················
AA 6153 hCoV-19/Saudi Arabia/KAUST-MADINAH1689/2020|EPI_IS ···················· ·A·················· ···················· ···················· ···················· ··A·················
AA 6154 hCoV-19/Saudi Arabia/KAUST-MADINAH1674/2020|EPI_IS ···················· ···················· ···················· ···················· ········A··········· ····················
AA 6155 hCoV-19/Saudi Arabia/KAUST-MADINAH1667/2020|EPI_IS ···················· ·A·················· ···················· ···················· ···················· ··A·················
AA 6156 hCoV-19/Saudi Arabia/KAUST-MADINAH1651/2020|EPI_IS ···················· ···················· ···················· ···················· ···················· ····················
AA 6157 hCoV-19/Saudi Arabia/KAUST-MADINAH1655/2020|EPI_IS ···················· ···················· ···················· ···················· ···················· ····················
AA 6158 hCoV-19/India/DL-IGIB11304644/2020|EPI_ISL_751280| ···················· ·A·················· ···················· ···················· ···················· ··A·················
AA 6159 hCoV-19/India/DL-IGIB11304769/2020|EPI_ISL_751281| ···················· ···················· ···················· ···················· ···················· ····················
AA 6160 hCoV-19/India/DL-IGIB11304885/2020|EPI_ISL_751282| ···················· ·A·················· ···················· ···················· ···················· ··A·················
AA 6161 hCoV-19/India/DL-IGIB11305017/2020|EPI_ISL_751283| ···················· ···················· ···················· ···················· ···················· ····················
AA 6162 hCoV-19/India/DL-IGIB11305034/2020|EPI_ISL_751284| ···················· ·A·················· ···················· ···················· ···················· ··A·················
AA 6163 hCoV-19/India/DL-IGIB11305072/2020|EPI_ISL_751285| ···················· ···················· ···················· ···················· ···················· ····················
AA 6164 hCoV-19/India/DL-IGIB11305086/2020|EPI_ISL_751286| ···················· ·A·················· ···················· ···················· ···················· ··A·················
AA 6165 hCoV-19/India/DL-IGIB11305231/2020|EPI_ISL_751287| ···················· ·A·················· ···················· ···················· ···················· ··A·················
AA 6166 hCoV-19/India/DL-IGIB11301835/2020|EPI_ISL_751288| ···················· ·A·················· ···················· ···················· ···················· ··A·················
AA 6167 hCoV-19/India/DL-IGIB11303573/2020|EPI_ISL_751289| ···················· ·A·················· ···················· ···················· ···················· ··A·················
AA 6168 hCoV-19/India/DL-IGIB11304423/2020|EPI_ISL_751290| ···················· ···················· ···················· ···················· ···················· ····················
AA 6169 hCoV-19/India/DL-IGIB11304453/2020|EPI_ISL_751291| ···················· ·A·················· ···················· ···················· ···················· ··A·················
AA 6170 hCoV-19/India/DL-IGIB11304570/2020|EPI_ISL_751292| ···················· ·N·················· ···················· ···················· ···················· ··R·················

| | | | | | | | | |
|---|---|---|---|---|---|---|---|---|
| AA | 6171 | hCoV-19/India/DL-IGIB11305020/2020\|EPI_ISL_751293\| | ···················· | ·A·················· | ···················· | ···················· | ···················· | ···A················ |
| AA | 6172 | hCoV-19/India/DL-IGIB11305044/2020\|EPI_ISL_751294\| | ···················· | ·R·················· | ···················· | ···················· | ···················· | ···A················ |
| AA | 6173 | hCoV-19/India/DL-IGIB11305164/2020\|EPI_ISL_751295\| | ···················· | ·A·················· | ···················· | ···················· | ···················· | ···A················ |
| AA | 6174 | hCoV-19/India/DL-IGIB11303588/2020\|EPI_ISL_751296\| | ···················· | ·A·················· | ···················· | ···················· | ···················· | ···A················ |
| AA | 6175 | hCoV-19/India/DL-IGIB11303760/2020\|EPI_ISL_751297\| | ···················· | ·A·················· | ···················· | ···················· | ···················· | ···A················ |
| AA | 6176 | hCoV-19/India/DL-IGIB11303963/2020\|EPI_ISL_751298\| | ···················· | ·A·················· | ···················· | ···················· | ···················· | ···A················ |
| AA | 6177 | hCoV-19/India/DL-IGIB11303980/2020\|EPI_ISL_751299\| | ···················· | ·A·················· | ···················· | ···················· | ···················· | ···A················ |
| AA | 6178 | hCoV-19/India/DL-IGIB11304239/2020\|EPI_ISL_751300\| | ···················· | ·A·················· | ···················· | ···················· | ···················· | ···A················ |
| AA | 6179 | hCoV-19/India/DL-IGIB11304386/2020\|EPI_ISL_751301\| | ···················· | ·A·················· | ···················· | ···················· | ···················· | ···A················ |
| AA | 6180 | hCoV-19/India/DL-IGIB11304392/2020\|EPI_ISL_751302\| | ···················· | ·A·················· | ···················· | ···················· | ···················· | ···A················ |
| AA | 6181 | hCoV-19/India/DL-IGIB11304400/2020\|EPI_ISL_751303\| | ···················· | ·A·················· | ···················· | ···················· | ···················· | ···A················ |
| AA | 6182 | hCoV-19/India/DL-IGIB11304669/2020\|EPI_ISL_751304\| | ···················· | ···················· | ···················· | ···················· | ···················· | ···················· |
| AA | 6183 | hCoV-19/India/DL-IGIB11304767/2020\|EPI_ISL_751305\| | ···················· | ·A·················· | ···················· | ···················· | ···················· | ···A················ |
| AA | 6184 | hCoV-19/India/DL-IGIB11304857/2020\|EPI_ISL_751306\| | ···················· | ·A·················· | ···················· | ···················· | ···················· | ···A················ |
| AA | 6185 | hCoV-19/India/DL-IGIB11305065/2020\|EPI_ISL_751307\| | ···················· | ·R·················· | ···················· | ···················· | ···················· | ···R················ |
| AA | 6186 | hCoV-19/India/DL-IGIB11305171/2020\|EPI_ISL_751308\| | ···················· | ·A·················· | ···················· | ···················· | ···················· | ···A················ |
| AA | 6187 | hCoV-19/India/DL-IGIB11305763/2020\|EPI_ISL_751309\| | ···················· | ·A·················· | ···················· | ···················· | ···················· | ···A················ |
| AA | 6188 | hCoV-19/India/DL-IGIB11305680/2020\|EPI_ISL_751311\| | ···················· | ·A·················· | ···················· | ···················· | ···················· | ···A················ |
| AA | 6189 | hCoV-19/India/DL-IGIB11305615/2020\|EPI_ISL_751312\| | ···················· | ·A·················· | ···················· | ···················· | ···················· | ···A················ |
| AA | 6190 | hCoV-19/Ireland/D-NVRL-78IRL71543/2020\|EPI_ISL_752 | ···················· | ···················· | ···················· | ···················· | ···················· | ···················· |
| AA | 6191 | hCoV-19/Ireland/MN-NVRL-78IRL71980/2020\|EPI_ISL_75 | ···················· | ···················· | ···················· | ···················· | ···················· | ···················· |
| AA | 6192 | hCoV-19/Ireland/TA-NVRL-78IRL73009/2020\|EPI_ISL_75 | ···················· | ···················· | ···················· | ···················· | ···················· | ···················· |
| AA | 6193 | hCoV-19/Ireland/WH-NVRL-78IRL73030/2020\|EPI_ISL_75 | ···················· | ···················· | ···················· | ···················· | ···················· | ···················· |
| AA | 6194 | hCoV-19/Ireland/KE-NVRL-78IRL61834/2020\|EPI_ISL_75 | ···················· | ···················· | ···················· | ···················· | ···················· | ···················· |
| AA | 6195 | hCoV-19/Ireland/LK-NVRL-78IRL69457/2020\|EPI_ISL_75 | ···················· | ···················· | ···················· | ···················· | ···················· | ···················· |
| AA | 6196 | hCoV-19/Ireland/KE-NVRL-78IRL70189/2020\|EPI_ISL_75 | ···················· | ···················· | ···················· | ···················· | ···················· | ···················· |
| AA | 6197 | hCoV-19/Ireland/WW-NVRL-78IRL70235/2020\|EPI_ISL_75 | ···················· | ···················· | ···················· | ···················· | ···················· | ···················· |
| AA | 6198 | hCoV-19/Ireland/CO-NVRL-78IRL70434/2020\|EPI_ISL_75 | ···················· | ···················· | ···················· | ···················· | ···················· | ···················· |
| AA | 6199 | hCoV-19/Ireland/G-NVRL-78IRL71368/2020\|EPI_ISL_752 | ···················· | ···················· | ···················· | ···················· | ···················· | ···················· |
| AA | 6200 | hCoV-19/Ireland/KE-NVRL-78IRL71660/2020\|EPI_ISL_75 | ···················· | ···················· | ···················· | ···················· | ···················· | ···················· |
| AA | 6201 | hCoV-19/Palestine/AAS25/2020\|EPI_ISL_752605\|2020-1 | ···················· | ···················· | ···················· | ···················· | ···················· | ···················· |
| AA | 6202 | hCoV-19/USA/HI-H200007/2020\|EPI_ISL_752608\|2020-03 | ···················· | ···················· | ···················· | ···················· | ···················· | ···················· |
| AA | 6203 | hCoV-19/USA/HI-H200011/2020\|EPI_ISL_752609\|2020-03 | ···················· | ···················· | ···················· | ···················· | ···················· | ···················· |
| AA | 6204 | hCoV-19/USA/HI-H200014/2020\|EPI_ISL_752610\|2020-04 | ···················· | ···················· | ···················· | ···················· | ···················· | ···················· |
| AA | 6205 | hCoV-19/USA/HI-H200015/2020\|EPI_ISL_752611\|2020-04 | ···················· | ···················· | ···················· | ···················· | ···················· | ···················· |
| AA | 6206 | hCoV-19/USA/HI-H200016/2020\|EPI_ISL_752612\|2020-03 | ···················· | ···················· | ···················· | ···················· | ···················· | ···················· |
| AA | 6207 | hCoV-19/USA/HI-H200019/2020\|EPI_ISL_752613\|2020-04 | ···················· | ···················· | ···················· | ···················· | ···················· | ···················· |
| AA | 6208 | hCoV-19/USA/HI-H200027/2020\|EPI_ISL_752614\|2020-03 | ···················· | ···················· | ···················· | ···················· | ···················· | ···················· |
| AA | 6209 | hCoV-19/USA/HI-H200028/2020\|EPI_ISL_752615\|2020-03 | ···················· | ···················· | ···················· | ···················· | ···················· | ···················· |
| AA | 6210 | hCoV-19/USA/HI-H200030/2020\|EPI_ISL_752616\|2020-03 | ···················· | ···················· | ···················· | ···················· | ···················· | ···················· |
| AA | 6211 | hCoV-19/USA/HI-H200031/2020\|EPI_ISL_752617\|2020-03 | ···················· | ···················· | ···················· | ···················· | ···················· | ···················· |
| AA | 6212 | hCoV-19/USA/HI-H200032/2020\|EPI_ISL_752618\|2020-03 | ···················· | ···················· | ···················· | ···················· | ···················· | ···················· |
| AA | 6213 | hCoV-19/USA/HI-H200034/2020\|EPI_ISL_752619\|2020-03 | ···················· | ···················· | ···················· | ···················· | ···················· | ···················· |
| AA | 6214 | hCoV-19/USA/HI-H200036/2020\|EPI_ISL_752620\|2020-04 | ···················· | ···················· | ···················· | ···················· | ···················· | ···················· |
| AA | 6215 | hCoV-19/USA/HI-H200037/2020\|EPI_ISL_752621\|2020-04 | ···················· | ···················· | ···················· | ···················· | ···················· | ···················· |
| AA | 6216 | hCoV-19/USA/HI-H200038/2020\|EPI_ISL_752622\|2020-04 | ···················· | ···················· | ···················· | ···················· | ···················· | ···················· |
| AA | 6217 | hCoV-19/USA/HI-H200039/2020\|EPI_ISL_752623\|2020-04 | ···················· | ···················· | ···················· | ···················· | ···················· | ···················· |
| AA | 6218 | hCoV-19/USA/HI-H200040/2020\|EPI_ISL_752624\|2020-04 | ···················· | ···················· | ···················· | ···················· | ···················· | ···················· |
| AA | 6219 | hCoV-19/USA/HI-H200041/2020\|EPI_ISL_752625\|2020-04 | ···················· | ···················· | ···················· | ···················· | ···················· | ···················· |
| AA | 6220 | hCoV-19/USA/HI-H200042/2020\|EPI_ISL_752626\|2020-03 | ···················· | ···················· | ···················· | ···················· | ···················· | ···················· |
| AA | 6221 | hCoV-19/USA/HI-H200043-1/2020\|EPI_ISL_752627\|2020- | ···················· | ···················· | ···················· | ···················· | ···················· | ···················· |
| AA | 6222 | hCoV-19/USA/HI-H200043-2/2020\|EPI_ISL_752628\|2020- | ···················· | ···················· | ···················· | ···················· | ···················· | ···················· |
| AA | 6223 | hCoV-19/USA/HI-H200044/2020\|EPI_ISL_752629\|2020-04 | ···················· | ···················· | ···················· | ···················· | ···················· | ···················· |
| AA | 6224 | hCoV-19/USA/HI-H200045/2020\|EPI_ISL_752630\|2020-04 | ···················· | ···················· | ···················· | ···················· | ···················· | ···················· |
| AA | 6225 | hCoV-19/USA/HI-H200046/2020\|EPI_ISL_752631\|2020-04 | ···················· | ···················· | ···················· | ···················· | ···················· | ···················· |
| AA | 6226 | hCoV-19/USA/HI-H200047/2020\|EPI_ISL_752632\|2020-04 | ···················· | ···················· | ···················· | ···················· | ···················· | ···················· |
| AA | 6227 | hCoV-19/USA/HI-H200048/2020\|EPI_ISL_752633\|2020-04 | ···················· | ···················· | ···················· | ···················· | ···················· | ···················· |
| AA | 6228 | hCoV-19/USA/HI-H200049/2020\|EPI_ISL_752634\|2020-04 | ···················· | ···················· | ···················· | ···················· | ···················· | ···················· |
| AA | 6229 | hCoV-19/USA/HI-H200050/2020\|EPI_ISL_752635\|2020-04 | ···················· | ···················· | ···················· | ···················· | ···················· | ···················· |
| AA | 6230 | hCoV-19/USA/HI-H200051/2020\|EPI_ISL_752636\|2020-04 | ···················· | ···················· | ···················· | ···················· | ···················· | ···················· |
| AA | 6231 | hCoV-19/USA/HI-H200052/2020\|EPI_ISL_752637\|2020-04 | ···················· | ···················· | ···················· | ···················· | ···················· | ···················· |
| AA | 6232 | hCoV-19/USA/HI-H200053/2020\|EPI_ISL_752638\|2020-04 | ···················· | ···················· | ···················· | ···················· | ···················· | ···················· |
| AA | 6233 | hCoV-19/USA/HI-H200054/2020\|EPI_ISL_752639\|2020-04 | ···················· | ···················· | ···················· | ···················· | ···················· | ···················· |
| AA | 6234 | hCoV-19/USA/HI-H200056/2020\|EPI_ISL_752641\|2020-04 | ···················· | ···················· | ···················· | ···················· | ···················· | ···················· |
| AA | 6235 | hCoV-19/USA/HI-H200057/2020\|EPI_ISL_752642\|2020-05 | ···················· | ···················· | ···················· | ···················· | ···················· | ···················· |
| AA | 6236 | hCoV-19/USA/HI-H200059/2020\|EPI_ISL_752643\|2020-04 | ···················· | ···················· | ···················· | ···················· | ···················· | ···················· |
| AA | 6237 | hCoV-19/Germany/DE-BE-ChVir-D1940-8248/2020\|EPI_IS | ···················· | ·A·················· | ···················· | ···················· | ···················· | ···················· |
| AA | 6238 | hCoV-19/Germany/DE-BE-ChVir-D1941-8263/2020\|EPI_IS | ···················· | ···················· | ···················· | ···················· | ···················· | ···················· |
| AA | 6239 | hCoV-19/Germany/DE-BE-ChVir-D1942-4496/2020\|EPI_IS | ···················· | ·A·················· | ···················· | ···················· | ···················· | ···················· |
| AA | 6240 | hCoV-19/Germany/DE-BE-ChVir-D1943-5032/2020\|EPI_IS | ···················· | ···················· | ···················· | ···················· | ···················· | ···················· |
| AA | 6241 | hCoV-19/Germany/DE-BE-ChVir-D1945-8218/2020\|EPI_IS | ···················· | ···················· | ···················· | ···················· | ···················· | ···················· |
| AA | 6242 | hCoV-19/Germany/DE-BE-ChVir-D1948-8295/2020\|EPI_IS | ···················· | ·A·················· | ···················· | ···················· | ···················· | ···················· |
| AA | 6243 | hCoV-19/Germany/DE-BE-ChVir-D1952-5748/2020\|EPI_IS | ···················· | ·A·················· | ···················· | ···················· | ···················· | ···················· |
| AA | 6244 | hCoV-19/Germany/DE-BE-ChVir-D1953-7492/2020\|EPI_IS | ···················· | ·A·················· | ···················· | ···················· | ···················· | ···················· |
| AA | 6245 | hCoV-19/Germany/DE-BE-ChVir-D1956-8255/2020\|EPI_IS | ···················· | ···················· | ···················· | ···················· | ···················· | ···················· |
| AA | 6246 | hCoV-19/Germany/DE-BE-ChVir-D1957-4437/2020\|EPI_IS | ···················· | ···················· | ···················· | ···················· | ······T············· | ···················· |
| AA | 6247 | hCoV-19/Germany/DE-BE-ChVir-D1958-8276/2020\|EPI_IS | ···················· | ···················· | ···················· | ···················· | ···················· | ···················· |
| AA | 6248 | hCoV-19/Germany/DE-BE-ChVir-D1959-4442/2020\|EPI_IS | ···················· | ···················· | ···················· | ···················· | ···················· | ···················· |
| AA | 6249 | hCoV-19/Germany/DE-BE-ChVir-D1996-4483/2020\|EPI_IS | ···················· | ···················· | ···················· | ···················· | ···················· | ···················· |
| AA | 6250 | hCoV-19/Germany/DE-BE-ChVir-D1997-8209/2020\|EPI_IS | ···················· | ·A·················· | ···················· | ···················· | ···················· | ···················· |

| | | | | | | | |
|---|---|---|---|---|---|---|---|
| AA | 6251 hCoV-19/Germany/DE-BE-ChVir-D1998-4426/2020\|EPI_IS | ···················· | ···················· | ···················· | ···················· | ···················· | ···················· |
| AA | 6252 hCoV-19/Germany/DE-BE-ChVir-D1999-8262/2020\|EPI_IS | ···················· | ···················· | ···················· | ···················· | ···················· | ···················· |
| AA | 6253 hCoV-19/Germany/DE-BE-ChVir-D2000-8271/2020\|EPI_IS | ···················· | ·A·················· | ···················· | ···················· | ···················· | ···················· |
| AA | 6254 hCoV-19/Germany/DE-BE-ChVir-D2003-5974/2020\|EPI_IS | ···················· | ···················· | ···················· | ···················· | ···················· | ···················· |
| AA | 6255 hCoV-19/Germany/DE-BE-ChVir-D2004-8247/2020\|EPI_IS | ···················· | ···················· | ···················· | ···················· | ······T············· | ···················· |
| AA | 6256 hCoV-19/Germany/DE-BE-ChVir-D2006-8256/2020\|EPI_IS | ···················· | ···················· | ···················· | ···················· | ···················· | ···················· |
| AA | 6257 hCoV-19/Germany/DE-BE-ChVir-D2007-8200/2020\|EPI_IS | ···················· | ···················· | ···················· | ···················· | ···················· | ···················· |
| AA | 6258 hCoV-19/Germany/DE-BE-ChVir-D2008-8215/2020\|EPI_IS | ···················· | ···················· | ···················· | ···················· | ···················· | ···················· |
| AA | 6259 hCoV-19/Germany/DE-BE-ChVir-W1074-8144/2020\|EPI_IS | ···················· | ···················· | ···················· | ···················· | ···················· | ···················· |
| AA | 6260 hCoV-19/Germany/DE-BE-ChVir-W1078-7878/2020\|EPI_IS | ···················· | ···················· | ···················· | ···················· | ···················· | ···················· |
| AA | 6261 hCoV-19/Germany/DE-BE-ChVir-W1081-8197/2020\|EPI_IS | ···················· | ···················· | ···················· | ···················· | ···················· | ···················· |
| AA | 6262 hCoV-19/Germany/DE-BE-ChVir-W1082-8115/2020\|EPI_IS | ···················· | ···················· | ···················· | ···················· | ···················· | ···················· |
| AA | 6263 hCoV-19/Germany/DE-BE-ChVir-W1084-8118/2020\|EPI_IS | ···················· | ···················· | ···················· | ···················· | ···················· | ···················· |
| AA | 6264 hCoV-19/Germany/DE-BE-ChVir-W1133-5742/2020\|EPI_IS | ···················· | ···················· | ···················· | ···················· | ···················· | ···················· |
| AA | 6265 hCoV-19/Germany/DE-BE-ChVir-W1134-8182/2020\|EPI_IS | ···················· | ···················· | ···················· | ···················· | ···················· | ···················· |
| AA | 6266 hCoV-19/Germany/DE-BE-ChVir-W1136-4353/2020\|EPI_IS | ···················· | ···················· | ···················· | ···················· | ···················· | ···················· |
| AA | 6267 hCoV-19/Germany/DE-BE-ChVir-W1137-5740/2020\|EPI_IS | ···················· | ···················· | ···················· | ···················· | ···················· | ···················· |
| AA | 6268 hCoV-19/Germany/DE-BE-ChVir-W1139-8123/2020\|EPI_IS | ···················· | ···················· | ···················· | ···················· | ···················· | ···················· |
| AA | 6269 hCoV-19/Germany/DE-BE-ChVir-W1140-8100/2020\|EPI_IS | ···················· | ···················· | ···················· | ···················· | ···················· | ···················· |
| AA | 6270 hCoV-19/Germany/DE-BE-ChVir-W1148-8141/2020\|EPI_IS | ···················· | ···················· | ···················· | ···················· | ···················· | ···················· |
| AA | 6271 hCoV-19/Germany/DE-BE-ChVir-W1149-8118/2020\|EPI_IS | ···················· | ···················· | ···················· | ···················· | ···················· | ···················· |
| AA | 6272 hCoV-19/Germany/DE-BE-ChVir-W1151-4300/2020\|EPI_IS | ···················· | ···················· | ···················· | ···················· | ···················· | ···················· |
| AA | 6273 hCoV-19/Germany/DE-BE-ChVir-W1178-8106/2020\|EPI_IS | ···················· | ···················· | ···················· | ···················· | ···················· | ···················· |
| AA | 6274 hCoV-19/Germany/DE-BE-ChVir-W1281-8144/2020\|EPI_IS | ···················· | ···················· | ···················· | ···················· | ···················· | ···················· |
| AA | 6275 hCoV-19/Germany/DE-BE-ChVir-W1282-4384/2020\|EPI_IS | ···················· | ···················· | ···················· | ···················· | ···················· | ···················· |
| AA | 6276 hCoV-19/Germany/DE-BE-ChVir-W1285-4367/2020\|EPI_IS | ···················· | ···················· | ···················· | ···················· | ···················· | ···················· |
| AA | 6277 hCoV-19/Germany/DE-BE-ChVir-W1291-8183/2020\|EPI_IS | ···················· | ···················· | ···················· | ···················· | ···················· | ···················· |
| AA | 6279 hCoV-19/Germany/DE-BE-ChVir-W1299-4302/2020\|EPI_IS | ···················· | ···················· | ···················· | ···················· | ···················· | ···················· |
| AA | 6280 hCoV-19/Germany/DE-BE-ChVir-W1303-8106/2020\|EPI_IS | ···················· | ···················· | ···················· | ···················· | ···················· | ···················· |
| AA | 6281 hCoV-19/Germany/DE-BE-ChVir-W1309-8155/2020\|EPI_IS | ···················· | ···················· | ···················· | ···················· | ···················· | ···················· |
| AA | 6282 hCoV-19/Germany/DE-BE-ChVir-W1336-5617/2020\|EPI_IS | ···················· | ···················· | ···················· | ···················· | ···················· | ···················· |
| AA | 6283 hCoV-19/Germany/DE-BE-ChVir-W1340-8291/2020\|EPI_IS | ···················· | ···················· | ···················· | ···················· | ···················· | ···················· |
| AA | 6284 hCoV-19/Germany/DE-BE-ChVir-W1414-5697/2020\|EPI_IS | ···················· | ···················· | ···················· | ···················· | ···················· | ···················· |
| AA | 6285 hCoV-19/Germany/DE-BE-ChVir-W1414-6707/2020\|EPI_IS | ···················· | ···················· | ···················· | ···················· | ···················· | ···················· |
| AA | 6286 hCoV-19/Germany/DE-BE-ChVir-W1416-4346/2020\|EPI_IS | ······C············· | ···················· | ···················· | ···················· | ···················· | ···················· |
| AA | 6287 hCoV-19/Germany/DE-BE-ChVir-W1417-8240/2020\|EPI_IS | ···················· | ···················· | ···················· | ···················· | ···················· | ···················· |
| AA | 6288 hCoV-19/Germany/DE-BE-ChVir-W1418-8203/2020\|EPI_IS | ···················· | ···················· | ···················· | ···················· | ···················· | ···················· |
| AA | 6289 hCoV-19/Germany/DE-BE-ChVir-W1427-8261/2020\|EPI_IS | ···················· | ···················· | ···················· | ···················· | ···················· | ···················· |
| AA | 6290 hCoV-19/Germany/DE-BE-ChVir-W1429-8283/2020\|EPI_IS | ···················· | ···················· | ···················· | ···················· | ···················· | ···················· |
| AA | 6291 hCoV-19/Germany/DE-BE-ChVir-W1431-8290/2020\|EPI_IS | ···················· | ···················· | ···················· | ···················· | ···················· | ···················· |
| AA | 6292 hCoV-19/Germany/DE-BE-ChVir-W1432-4374/2020\|EPI_IS | ···················· | ···················· | ···················· | ···················· | ···················· | ···················· |
| AA | 6293 hCoV-19/Germany/DE-BE-ChVir-W1433-8258/2020\|EPI_IS | ···················· | ···················· | ···················· | ···················· | ···················· | ···················· |
| AA | 6294 hCoV-19/Germany/DE-BE-ChVir-W1436-4360/2020\|EPI_IS | ···················· | ···················· | ···················· | ···················· | ···················· | ···················· |
| AA | 6295 hCoV-19/Germany/DE-BE-ChVir-W1439-5711/2020\|EPI_IS | ···················· | ···················· | ···················· | ···················· | ···················· | ···················· |
| AA | 6296 hCoV-19/Germany/DE-BE-ChVir-W1440-8273/2020\|EPI_IS | ···················· | ···················· | ···················· | ···················· | ···················· | ···················· |
| AA | 6297 hCoV-19/Germany/DE-BE-ChVir-W1443-8219/2020\|EPI_IS | ···················· | ···················· | ···················· | ···················· | ···················· | ···················· |
| AA | 6298 hCoV-19/Germany/DE-BE-ChVir-W1449-8226/2020\|EPI_IS | ···················· | ···················· | ···················· | ···················· | ···················· | ···················· |
| AA | 6299 hCoV-19/Germany/DE-BE-ChVir-W1451-8252/2020\|EPI_IS | ···················· | ···················· | ···················· | ···················· | ···················· | ···················· |
| AA | 6300 hCoV-19/Germany/DE-BE-ChVir-W1635-6790/2020\|EPI_IS | ···················· | ·A·················· | ···················· | ···················· | ···················· | ···················· |
| AA | 6301 hCoV-19/Germany/DE-BE-ChVir-W1637-6714/2020\|EPI_IS | ···················· | ···················· | ···················· | ···················· | ···················· | ···················· |
| AA | 6302 hCoV-19/Finland/3LS18AG4/2020\|EPI_ISL_756100\|2020- | ···················· | ·A·················· | ···················· | ···················· | ···················· | ···A················ |
| AA | 6303 hCoV-19/Finland/3LS20H5/2020\|EPI_ISL_756101\|2020-1 | ···················· | ···················· | ···················· | ···················· | ···················· | ···················· |
| AA | 6304 hCoV-19/Finland/3LS22E11/2020\|EPI_ISL_756102\|2020- | ···················· | ···················· | ···················· | ···················· | ···················· | ···················· |
| AA | 6305 hCoV-19/Finland/3LS2AD11/2020\|EPI_ISL_756103\|2020- | ···················· | ···················· | ···················· | ···················· | ···················· | ···················· |
| AA | 6306 hCoV-19/Finland/3LS2AF5/2020\|EPI_ISL_756104\|2020-1 | ···················· | ·A·················· | ···················· | ···················· | ···················· | ···A················ |
| AA | 6307 hCoV-19/Finland/3LS3AA4/2020\|EPI_ISL_756105\|2020-1 | ···················· | ·A·················· | ···················· | ···················· | ···················· | ···A················ |
| AA | 6308 hCoV-19/Finland/3LS3AB5/2020\|EPI_ISL_756106\|2020-1 | ···················· | ···················· | ···················· | ···················· | ···················· | ···················· |
| AA | 6309 hCoV-19/Finland/3LS5A11/2020\|EPI_ISL_756107\|2020-1 | ···················· | ·A·················· | ···················· | ···················· | ···················· | ···A················ |
| AA | 6310 hCoV-19/Finland/3LS5B11/2020\|EPI_ISL_756108\|2020-1 | ···················· | ·A·················· | ···················· | ···················· | ···················· | ···A················ |
| AA | 6311 hCoV-19/Finland/3LS5BF1/2020\|EPI_ISL_756109\|2020-1 | ···················· | ·A·················· | ···················· | ···················· | ···················· | ···A················ |
| AA | 6312 hCoV-19/Finland/3LS6E7/2020\|EPI_ISL_756110\|2020-10 | ···················· | ·A·················· | ···················· | ···················· | ···················· | ···A················ |
| AA | 6313 hCoV-19/Finland/3LS7BD7/2020\|EPI_ISL_756111\|2020-1 | ···················· | ·A·················· | ···················· | ···················· | ···················· | ···A················ |
| AA | 6314 hCoV-19/Finland/2LS12D1/2020\|EPI_ISL_756112\|2020-1 | ···················· | ···················· | ···················· | ···················· | ···················· | ···················· |
| AA | 6315 hCoV-19/Finland/2LS12G2/2020\|EPI_ISL_756113\|2020-1 | ···················· | ···················· | ···················· | ···················· | ···················· | ···················· |
| AA | 6316 hCoV-19/Finland/2LS13A11/2020\|EPI_ISL_756114\|2020- | ···················· | ···················· | ···················· | ···················· | ···················· | ···················· |
| AA | 6317 hCoV-19/Finland/2LS13A5/2020\|EPI_ISL_756115\|2020-1 | ···················· | ···················· | ···················· | ···················· | ···················· | ···················· |
| AA | 6318 hCoV-19/Finland/2LS13A7/2020\|EPI_ISL_756116\|2020-1 | ···················· | ···················· | ···················· | ···················· | ···················· | ···················· |
| AA | 6319 hCoV-19/Finland/2LS13H9/2020\|EPI_ISL_756117\|2020-1 | ···················· | ···················· | ···················· | ···················· | ···················· | ···················· |
| AA | 6320 hCoV-19/Finland/2LS16E1/2020\|EPI_ISL_756118\|2020-1 | ···················· | ·A·················· | ···················· | ···················· | ···················· | ···A················ |
| AA | 6321 hCoV-19/Finland/2LS18C11/2020\|EPI_ISL_756119\|2020- | ···················· | ···················· | ···················· | ···················· | ···················· | ···················· |
| AA | 6322 hCoV-19/Finland/2LS2F1/2020\|EPI_ISL_756120\|2020-10 | ···················· | ·A·················· | ···················· | ···················· | ···················· | ···A················ |
| AA | 6323 hCoV-19/Finland/2LS3B11/2020\|EPI_ISL_756121\|2020-1 | ···················· | ·A·················· | ···················· | ···················· | ···················· | ···A················ |
| AA | 6324 hCoV-19/Finland/2LS5A3/2020\|EPI_ISL_756122\|2020-10 | ···················· | ···················· | ···················· | ···················· | ···················· | ···················· |
| AA | 6325 hCoV-19/Finland/2LS5E11/2020\|EPI_ISL_756123\|2020-1 | ···················· | ···················· | ···················· | ···················· | ···················· | ···················· |
| AA | 6326 hCoV-19/Finland/2LS7A3/2020\|EPI_ISL_756124\|2020-10 | ···················· | ·A·················· | ···················· | ···················· | ···················· | ···A················ |
| AA | 6327 hCoV-19/Finland/2LS9F6/2020\|EPI_ISL_756125\|2020-10 | ···················· | ·A·················· | ···················· | ···················· | ···················· | ···A················ |
| AA | 6328 hCoV-19/USA/WY-WPHL-20156925/2020\|EPI_ISL_756126\|2 | ···················· | ···················· | ···················· | ···················· | ···················· | ···················· |
| AA | 6329 hCoV-19/USA/WY-WPHL-20157033/2020\|EPI_ISL_756128\|2 | ···················· | ···················· | ···················· | ···················· | ···················· | ···················· |
| AA | 6330 hCoV-19/USA/WY-WPHL-20157050/2020\|EPI_ISL_756129\|2 | ···················· | ···················· | ···················· | ···················· | ···················· | ···················· |
| AA | 6331 hCoV-19/USA/WY-WPHL-20157057/2020\|EPI_ISL_756130\|2 | ···················· | ···················· | ···················· | ···················· | ···················· | ···················· |

| | | | | | | | |
|---|---|---|---|---|---|---|---|
| AA | 6332 hCoV-19/USA/WY-WPHL-20157060/2020\|EPI_ISL_756131\|2 | ···················· | ···················· | ···················· | ···················· | ···················· | ···················· |
| AA | 6333 hCoV-19/USA/WY-WPHL-20157068/2020\|EPI_ISL_756132\|2 | ···················· | ···················· | ···················· | ···················· | ···················· | ···················· |
| AA | 6334 hCoV-19/USA/WY-WPHL-20157073/2020\|EPI_ISL_756133\|2 | ···················· | ·A·················· | ···················· | ···················· | ···················· | ···················· |
| AA | 6335 hCoV-19/USA/WY-WPHL-20157098/2020\|EPI_ISL_756134\|2 | ···················· | ···················· | ···················· | ···················· | ···················· | ···················· |
| AA | 6336 hCoV-19/England/205141162/2020\|EPI_ISL_756136\|2020 | ···················· | ···················· | ···················· | ···················· | ···················· | ···················· |
| AA | 6337 hCoV-19/England/205141163/2020\|EPI_ISL_756137\|2020 | ···················· | ···················· | ···················· | ···················· | ······T············· | ···················· |
| AA | 6338 hCoV-19/England/205141167/2020\|EPI_ISL_756138\|2020 | ···················· | ···················· | ···················· | ···················· | ···················· | ···················· |
| AA | 6340 hCoV-19/England/205141175/2020\|EPI_ISL_756140\|2020 | ···················· | ···················· | ···················· | ···················· | ··N··N·············· | ···················· |
| AA | 6342 hCoV-19/England/205141183/2020\|EPI_ISL_756142\|2020 | ···················· | ···················· | ···················· | ···················· | ···················· | ···················· |
| AA | 6343 hCoV-19/England/205141189/2020\|EPI_ISL_756143\|2020 | ···················· | ···················· | ···················· | ···················· | ···················· | ···················· |
| AA | 6345 hCoV-19/England/205141198/2020\|EPI_ISL_756146\|2020 | ···················· | ···················· | ···················· | ···················· | ···················· | ···················· |
| AA | 6346 hCoV-19/England/205141207/2020\|EPI_ISL_756150\|2020 | ···················· | ···················· | ···················· | ···················· | ···················· | ···················· |
| AA | 6347 hCoV-19/England/205141213/2020\|EPI_ISL_756152\|2020 | ···················· | ···················· | ···················· | ···················· | ···················· | ···················· |
| AA | 6348 hCoV-19/England/205141214/2020\|EPI_ISL_756153\|2020 | ···················· | ···················· | ···················· | ···················· | ···················· | ···················· |
| AA | 6349 hCoV-19/England/205141218/2020\|EPI_ISL_756155\|2020 | ···················· | ···················· | ···················· | ···················· | ···················· | ···················· |
| AA | 6350 hCoV-19/England/205141222/2020\|EPI_ISL_756157\|2020 | ···················· | ···················· | ···················· | ···················· | ···················· | ···················· |
| AA | 6351 hCoV-19/England/205141224/2020\|EPI_ISL_756158\|2020 | ···················· | ···················· | ···················· | ···················· | ···················· | ···················· |
| AA | 6352 hCoV-19/England/205141228/2020\|EPI_ISL_756159\|2020 | ···················· | ···················· | ···················· | ···················· | ···················· | ···················· |
| AA | 6353 hCoV-19/England/205141230/2020\|EPI_ISL_756160\|2020 | ···················· | ···················· | ···················· | ···················· | ···················· | ···················· |
| AA | 6354 hCoV-19/England/205141234/2020\|EPI_ISL_756161\|2020 | ···················· | ···················· | ···················· | ···················· | ···················· | ···················· |
| AA | 6355 hCoV-19/USA/WY-WPHL-20157230/2020\|EPI_ISL_756164\|2 | ···················· | ···················· | ···················· | ···················· | ···················· | ···················· |
| AA | 6356 hCoV-19/Finland/15ES15BE1/2020\|EPI_ISL_756165\|2020 | ···················· | ·A·················· | ···················· | ···················· | ···················· | ···A················ |
| AA | 6357 hCoV-19/Finland/15ES7C1/2020\|EPI_ISL_756166\|2020-0 | ···················· | ···················· | ···················· | ···················· | ···················· | ···················· |
| AA | 6358 hCoV-19/Finland/15LS14AA11/2020\|EPI_ISL_756167\|202 | ···················· | ·A·················· | ···················· | ···················· | ···················· | ···A················ |
| AA | 6359 hCoV-19/Finland/15LS14AB1/2020\|EPI_ISL_756168\|2020 | ···················· | ···················· | ···················· | ···················· | ···················· | ···················· |
| AA | 6360 hCoV-19/Finland/15LS14AE7/2020\|EPI_ISL_756169\|2020 | ···················· | ···················· | ···················· | ···················· | ···················· | ···················· |
| AA | 6361 hCoV-19/Denmark/DCGC-22432/2020\|EPI_ISL_757452\|202 | ···················· | ···················· | ···················· | ···················· | ···················· | ···················· |
| AA | 6362 hCoV-19/Denmark/DCGC-22473/2020\|EPI_ISL_757453\|202 | ···················· | ···················· | ···················· | ···················· | ···················· | ···················· |
| AA | 6363 hCoV-19/Denmark/DCGC-22512/2020\|EPI_ISL_757454\|202 | ···················· | ···················· | ···················· | ···················· | ···················· | ···················· |
| AA | 6364 hCoV-19/Denmark/DCGC-22541/2020\|EPI_ISL_757455\|202 | ···················· | ···················· | ···················· | ···················· | ···················· | ···················· |
| AA | 6365 hCoV-19/Denmark/DCGC-22556/2020\|EPI_ISL_757456\|202 | ···················· | ···················· | ···················· | ···················· | ···················· | ···················· |
| AA | 6366 hCoV-19/Denmark/DCGC-22562/2020\|EPI_ISL_757457\|202 | ···················· | ···················· | ···················· | ···················· | ···················· | ···················· |
| AA | 6367 hCoV-19/Denmark/DCGC-22581/2020\|EPI_ISL_757458\|202 | ···················· | ···················· | ···················· | ···················· | ···················· | ···················· |
| AA | 6368 hCoV-19/Denmark/DCGC-22591/2020\|EPI_ISL_757459\|202 | ···················· | ···················· | ···················· | ···················· | ···················· | ···················· |
| AA | 6369 hCoV-19/Denmark/DCGC-22619/2020\|EPI_ISL_757460\|202 | ···················· | ···················· | ···················· | ···················· | ···················· | ···················· |
| AA | 6370 hCoV-19/Denmark/DCGC-22630/2020\|EPI_ISL_757461\|202 | ···················· | ···················· | ···················· | ···················· | ···················· | ···················· |
| AA | 6371 hCoV-19/Denmark/DCGC-22583/2020\|EPI_ISL_757462\|202 | ···················· | ···················· | ···················· | ···················· | ···················· | ···················· |
| AA | 6372 hCoV-19/Denmark/DCGC-22391/2020\|EPI_ISL_757465\|202 | ···················· | ···················· | ···················· | ···················· | ···················· | ···················· |
| AA | 6373 hCoV-19/Denmark/DCGC-22395/2020\|EPI_ISL_757466\|202 | ···················· | ···················· | ···················· | ···················· | ···················· | ···················· |
| AA | 6374 hCoV-19/Denmark/DCGC-22424/2020\|EPI_ISL_757467\|202 | ···················· | ···················· | ···················· | ···················· | ···················· | ···················· |
| AA | 6375 hCoV-19/Denmark/DCGC-22483/2020\|EPI_ISL_757468\|202 | ···················· | ···················· | ···················· | ···················· | ···················· | ···················· |
| AA | 6376 hCoV-19/Denmark/DCGC-22608/2020\|EPI_ISL_757469\|202 | ···················· | ···················· | ···················· | ···················· | ···················· | ···················· |
| AA | 6377 hCoV-19/Denmark/DCGC-22381/2020\|EPI_ISL_757471\|202 | ···················· | ···················· | ···················· | ···················· | ···················· | ···················· |
| AA | 6378 hCoV-19/Denmark/DCGC-22387/2020\|EPI_ISL_757472\|202 | ···················· | ···················· | ···················· | ···················· | ···················· | ···················· |
| AA | 6379 hCoV-19/Denmark/DCGC-22407/2020\|EPI_ISL_757473\|202 | ···················· | ··G················· | ···················· | ···················· | ···················· | ···················· |
| AA | 6380 hCoV-19/Denmark/DCGC-22416/2020\|EPI_ISL_757474\|202 | ···················· | ···················· | ···················· | ···················· | ···················· | ···················· |
| AA | 6381 hCoV-19/Denmark/DCGC-22430/2020\|EPI_ISL_757475\|202 | ···················· | ···················· | ···················· | ···················· | ···················· | ···················· |
| AA | 6382 hCoV-19/Denmark/DCGC-22435/2020\|EPI_ISL_757476\|202 | ···················· | ···················· | ···················· | ···················· | ···················· | ···················· |
| AA | 6383 hCoV-19/Denmark/DCGC-22465/2020\|EPI_ISL_757477\|202 | ···················· | ···················· | ···················· | ···················· | ···················· | ···················· |
| AA | 6384 hCoV-19/Denmark/DCGC-22491/2020\|EPI_ISL_757478\|202 | ···················· | ···················· | ···················· | ···················· | ···················· | ···················· |
| AA | 6385 hCoV-19/Denmark/DCGC-22494/2020\|EPI_ISL_757479\|202 | ···················· | ···················· | ···················· | ···················· | ···················· | ···················· |
| AA | 6386 hCoV-19/Denmark/DCGC-22513/2020\|EPI_ISL_757480\|202 | ···················· | ···················· | ···················· | ···················· | ···················· | ···················· |
| AA | 6387 hCoV-19/Denmark/DCGC-22523/2020\|EPI_ISL_757481\|202 | ···················· | ···················· | ···················· | ···················· | ···················· | ···················· |
| AA | 6388 hCoV-19/Denmark/DCGC-22544/2020\|EPI_ISL_757482\|202 | ···················· | ···················· | ···················· | ···················· | ···················· | ···················· |
| AA | 6389 hCoV-19/Denmark/DCGC-22545/2020\|EPI_ISL_757483\|202 | ···················· | ···················· | ···················· | ···················· | ···················· | ···················· |
| AA | 6390 hCoV-19/Denmark/DCGC-22551/2020\|EPI_ISL_757484\|202 | ···················· | ···················· | ···················· | ···················· | ···················· | ···················· |
| AA | 6391 hCoV-19/Denmark/DCGC-22612/2020\|EPI_ISL_757485\|202 | ···················· | ···················· | ···················· | ···················· | ···················· | ···················· |
| AA | 6392 hCoV-19/Denmark/DCGC-22613/2020\|EPI_ISL_757486\|202 | ···················· | ···················· | ···················· | ···················· | ···················· | ···················· |
| AA | 6393 hCoV-19/Denmark/DCGC-22621/2020\|EPI_ISL_757487\|202 | ···················· | ···················· | ···················· | ···················· | ···················· | ···················· |
| AA | 6394 hCoV-19/Denmark/DCGC-22623/2020\|EPI_ISL_757488\|202 | ···················· | ···················· | ···················· | ···················· | ···················· | ···················· |
| AA | 6395 hCoV-19/Denmark/DCGC-22638/2020\|EPI_ISL_757489\|202 | ···················· | ···················· | ···················· | ···················· | ···················· | ···················· |
| AA | 6396 hCoV-19/Denmark/DCGC-22640/2020\|EPI_ISL_757490\|202 | ···················· | ···················· | ···················· | ···················· | ···················· | ···················· |
| AA | 6397 hCoV-19/Denmark/DCGC-22652/2020\|EPI_ISL_757491\|202 | ···················· | ···················· | ···················· | ···················· | ···················· | ···················· |
| AA | 6398 hCoV-19/Denmark/DCGC-22414/2020\|EPI_ISL_757494\|202 | ···················· | ···················· | ···················· | ···················· | ···················· | ···················· |
| AA | 6399 hCoV-19/Denmark/DCGC-22609/2020\|EPI_ISL_757495\|202 | ···················· | ·A·················· | ···················· | ···················· | ···················· | ···A················ |
| AA | 6400 hCoV-19/Denmark/DCGC-22357/2020\|EPI_ISL_758700\|202 | ···················· | ···················· | ···················· | ···················· | ···················· | ···················· |
| AA | 6401 hCoV-19/Denmark/DCGC-18342/2020\|EPI_ISL_749901\|202 | ···················· | ···················· | ···················· | ···················· | ···················· | ···················· |
| AA | 6402 hCoV-19/Denmark/DCGC-18343/2020\|EPI_ISL_749902\|202 | ···················· | ···················· | ···················· | ···················· | ···················· | ···················· |
| AA | 6403 hCoV-19/Denmark/DCGC-18346/2020\|EPI_ISL_749905\|202 | ···················· | ···················· | ···················· | ···················· | ···················· | ···················· |
| AA | 6404 hCoV-19/Denmark/DCGC-18347/2020\|EPI_ISL_749907\|202 | ···················· | ···················· | ···················· | ···················· | ···················· | ···················· |
| AA | 6405 hCoV-19/Denmark/DCGC-18348/2020\|EPI_ISL_749908\|202 | ···················· | ···················· | ···················· | ···················· | ···················· | ···················· |
| AA | 6406 hCoV-19/Denmark/DCGC-18349/2020\|EPI_ISL_749909\|202 | ···················· | ···················· | ···················· | ···················· | ···················· | ···················· |
| AA | 6407 hCoV-19/Denmark/DCGC-18350/2020\|EPI_ISL_749910\|202 | ···················· | ···················· | ···················· | ···················· | ···················· | ···················· |
| AA | 6408 hCoV-19/Denmark/DCGC-18351/2020\|EPI_ISL_749911\|202 | ···················· | ···················· | ···················· | ···················· | ···················· | ···················· |
| AA | 6409 hCoV-19/Denmark/DCGC-18352/2020\|EPI_ISL_749912\|202 | ···················· | ···················· | ···················· | ···················· | ···················· | ···················· |
| AA | 6410 hCoV-19/Denmark/DCGC-18354/2020\|EPI_ISL_749913\|202 | ···················· | ···················· | ···················· | ···················· | ···················· | ···················· |
| AA | 6411 hCoV-19/Denmark/DCGC-18355/2020\|EPI_ISL_749914\|202 | ···················· | ···················· | ···················· | ···················· | ···················· | ···················· |
| AA | 6412 hCoV-19/Denmark/DCGC-18356/2020\|EPI_ISL_749915\|202 | ···················· | ·A·················· | ···················· | ···················· | ···················· | ···A················ |
| AA | 6413 hCoV-19/India/DL-IGIB11305764/2020\|EPI_ISL_751310\| | ···················· | ·A·················· | ···················· | ···················· | ···················· | ···A················ |
| AA | 6414 hCoV-19/USA/HI-H200055/2020\|EPI_ISL_752640\|2020-04 | ···················· | ···················· | ···················· | ···················· | ···················· | ···················· |

AA 6415 hCoV-19/USA/HI-H200573/2020|EPI_ISL_753100|2020-09 ···················· ·A·················· ···················· ···················· ···················· ····················
AA 6416 hCoV-19/USA/HI-H200574/2020|EPI_ISL_753101|2020-09 ···················· ·A·················· ···················· ···················· ···················· ····················
AA 6417 hCoV-19/USA/HI-H200575/2020|EPI_ISL_753102|2020-09 ···················· ·A·················· ···················· ···················· ···················· ····················
AA 6418 hCoV-19/USA/HI-H200576/2020|EPI_ISL_753103|2020-09 ···················· ·A·················· ···················· ···················· ···················· ····················
AA 6419 hCoV-19/USA/HI-H200577/2020|EPI_ISL_753104|2020-09 ···················· ·A·················· ···················· ···················· ···················· ····················
AA 6420 hCoV-19/USA/HI-H200578/2020|EPI_ISL_753105|2020-10 ···················· ·A·················· ···················· ···················· ···················· ····················
AA 6421 hCoV-19/USA/HI-H200613/2020|EPI_ISL_753139|2020-11 ···················· ·A·················· ···················· ···················· ···················· ····················
AA 6422 hCoV-19/USA/HI-H200614/2020|EPI_ISL_753140|2020-11 ···················· ·A·················· ···················· ···················· ···················· ····················
AA 6423 hCoV-19/USA/HI-H200615/2020|EPI_ISL_753141|2020-11 ···················· ·A·················· ···················· ···················· ···················· ····················
AA 6424 hCoV-19/USA/HI-H200616/2020|EPI_ISL_753142|2020-11 ···················· ·A·················· ···················· ···················· ···················· ····················
AA 6425 hCoV-19/USA/HI-H200617/2020|EPI_ISL_753143|2020-11 ···················· ·A·················· ···················· ···················· ···················· ····················
AA 6426 hCoV-19/USA/HI-H200618/2020|EPI_ISL_753144|2020-11 ···················· ·A·················· ···················· ···················· ···················· ····················
AA 6427 hCoV-19/USA/HI-H200619/2020|EPI_ISL_753145|2020-11 ···················· ·A·················· ···················· ···················· ···················· ····················
AA 6428 hCoV-19/USA/HI-H200620/2020|EPI_ISL_753146|2020-11 ···················· ·A·················· ···················· ···················· ···················· ····················
AA 6429 hCoV-19/USA/HI-H200621/2020|EPI_ISL_753147|2020-09 ···················· ·A·················· ···················· ···················· ···················· ····················
AA 6430 hCoV-19/USA/HI-H200622/2020|EPI_ISL_753148|2020-09 ···················· ·A·················· ···················· ···················· ···················· ····················
AA 6431 hCoV-19/USA/HI-H200623/2020|EPI_ISL_753149|2020-09 ···················· ·A·················· ···················· ···················· ···················· ····················
AA 6432 hCoV-19/USA/HI-H200625/2020|EPI_ISL_753150|2020-11 ···················· ·A·················· ···················· ···················· ···················· ····················
AA 6433 hCoV-19/USA/HI-H200626/2020|EPI_ISL_753151|2020-11 ···················· ···················· ···················· ···················· ···················· ····················
AA 6434 hCoV-19/USA/HI-H200627/2020|EPI_ISL_753152|2020-11 ···················· ···················· ···················· ···················· ···················· ····················
AA 6435 hCoV-19/USA/HI-H200628/2020|EPI_ISL_753153|2020-11 ···················· ···················· ···················· ···················· ···················· ····················
AA 6436 hCoV-19/USA/HI-H200629/2020|EPI_ISL_753154|2020-11 ···················· ·A·················· ···················· ···················· ···················· ····················
AA 6437 hCoV-19/USA/HI-H200630/2020|EPI_ISL_753155|2020-11 ···················· ·A·················· ···················· ···················· ···················· ····················
AA 6438 hCoV-19/USA/HI-H200631/2020|EPI_ISL_753156|2020-11 ···················· ·A·················· ···················· ···················· ···················· ····················
AA 6439 hCoV-19/USA/HI-H200633/2020|EPI_ISL_753157|2020-11 ···················· ·A·················· ···················· ···················· ···················· ····················
AA 6440 hCoV-19/USA/HI-H200634/2020|EPI_ISL_753158|2020-11 ···················· ·A·················· ···················· ···················· ···················· ····················
AA 6441 hCoV-19/USA/HI-H200635/2020|EPI_ISL_753159|2020-11 ···················· ·A·················· ···················· ···················· ···················· ····················
AA 6442 hCoV-19/USA/HI-H200636/2020|EPI_ISL_753160|2020-11 ···················· ·A·················· ···················· ···················· ···················· ····················
AA 6443 hCoV-19/USA/HI-H200637/2020|EPI_ISL_753161|2020-11 ···················· ·A·················· ···················· ···················· ···················· ····················
AA 6444 hCoV-19/USA/HI-H200638/2020|EPI_ISL_753162|2020-11 ···················· ·A·················· ···················· ···················· ···················· ····················
AA 6445 hCoV-19/USA/HI-H200639/2020|EPI_ISL_753163|2020-11 ···················· ·A·················· ···················· ···················· ···················· ····················
AA 6446 hCoV-19/USA/HI-H200640/2020|EPI_ISL_753164|2020-11 ···················· ·A·················· ···················· ···················· ···················· ····················
AA 6447 hCoV-19/USA/HI-H200641/2020|EPI_ISL_753165|2020-11 ···················· ·A·················· ···················· ···················· ···················· ····················
AA 6448 hCoV-19/USA/HI-H200642/2020|EPI_ISL_753166|2020-11 ···················· ·A·················· ···················· ···················· ···················· ····················
AA 6449 hCoV-19/USA/HI-H200643/2020|EPI_ISL_753167|2020-11 ···················· ·A·················· ···················· ···················· ···················· ····················
AA 6450 hCoV-19/USA/HI-H200644/2020|EPI_ISL_753168|2020-11 ···················· ···················· ···················· ···················· ···················· ····················
AA 6451 hCoV-19/USA/HI-H200645/2020|EPI_ISL_753169|2020-11 ···················· ·A·················· ···················· ···················· ···················· ····················
AA 6452 hCoV-19/USA/HI-H200646/2020|EPI_ISL_753170|2020-11 ···················· ·A·················· ···················· ···················· ···················· ····················
AA 6453 hCoV-19/USA/HI-H200647/2020|EPI_ISL_753171|2020-11 ···················· ·A·················· ···················· ···················· ···················· ····················
AA 6454 hCoV-19/USA/HI-H200648/2020|EPI_ISL_753172|2020-11 ···················· ···················· ···················· ···················· ···················· ····················
AA 6455 hCoV-19/USA/HI-H200649/2020|EPI_ISL_753173|2020-11 ···················· ···················· ···················· ···················· ···················· ····················
AA 6456 hCoV-19/USA/HI-H200651/2020|EPI_ISL_753174|2020-11 ···················· ·A·················· ···················· ···················· ···················· ····················
AA 6457 hCoV-19/USA/HI-H200652/2020|EPI_ISL_753175|2020-11 ···················· ·A·················· ···················· ···················· ···················· ····················
AA 6458 hCoV-19/USA/HI-H200653/2020|EPI_ISL_753176|2020-11 ···················· ···················· ···················· ···················· ···················· ····················
AA 6459 hCoV-19/USA/HI-H200654/2020|EPI_ISL_753177|2020-11 ···················· ···················· ···················· ···················· ···················· ····················
AA 6460 hCoV-19/USA/HI-H200655/2020|EPI_ISL_753178|2020-12 ···················· ·A·················· ···················· ···················· ···················· ····················
AA 6461 hCoV-19/USA/HI-H200656/2020|EPI_ISL_753179|2020-12 ···················· ·A·················· ···················· ···················· ···················· ····················
AA 6462 hCoV-19/Germany/DE-BE-ChVir-D2012-8249/2020|EPI_IS ···················· ···················· ···················· ···················· ···················· ····················
AA 6463 hCoV-19/Germany/DE-BE-ChVir-D2016-5933/2020|EPI_IS ···················· ···················· ···················· ···················· ···················· ····················
AA 6464 hCoV-19/Germany/DE-BE-ChVir-D2017-8214/2020|EPI_IS ···················· ···A················ ···················· ···················· ···················· ····················
AA 6465 hCoV-19/Germany/DE-BE-ChVir-D2020-4418/2020|EPI_IS ···················· ···················· ···················· ···················· ···················· ····················
AA 6466 hCoV-19/Germany/DE-BE-ChVir-D2021-4462/2020|EPI_IS ···················· ···················· ···················· ···················· ···················· ····················
AA 6467 hCoV-19/Germany/DE-BE-ChVir-D2022-4412/2020|EPI_IS ···················· ···················· ···················· ···················· ···················· ····················
AA 6468 hCoV-19/Germany/DE-BE-ChVir-D2023-4491/2020|EPI_IS ···················· ···················· ···················· ···················· ···················· ····················
AA 6469 hCoV-19/Germany/DE-BE-ChVir-D2024-8294/2020|EPI_IS ···················· ···················· ···················· ···················· ···················· ····················
AA 6470 hCoV-19/Germany/DE-BE-ChVir-D2025-5941/2020|EPI_IS ···················· ···················· ···················· ···················· ···················· ····················
AA 6471 hCoV-19/Germany/DE-BE-ChVir-D2026-8257/2020|EPI_IS ···················· ···················· ···················· ···················· ···················· ····················
AA 6472 hCoV-19/Germany/DE-BE-ChVir-D2027-8242/2020|EPI_IS ···················· ···················· ···················· ···················· ···················· ····················
AA 6473 hCoV-19/Germany/DE-BE-ChVir-D2033-5902/2020|EPI_IS ···················· ·A·················· ···················· ···················· ···················· ····················
AA 6474 hCoV-19/Germany/DE-BE-ChVir-D2034-8234/2020|EPI_IS ···················· ···················· ···················· ···················· ···················· ····················
AA 6475 hCoV-19/Germany/DE-BE-ChVir-D2035-8263/2020|EPI_IS ···················· ···················· ···················· ···················· ···················· ····················
AA 6476 hCoV-19/Germany/DE-BE-ChVir-D2036-8292/2020|EPI_IS ···················· ···················· ···················· ···················· ···················· ····················
AA 6477 hCoV-19/Germany/DE-BE-ChVir-D2037-8226/2020|EPI_IS ···················· ···················· ···················· ···················· ···················· ····················
AA 6478 hCoV-19/Germany/DE-BE-ChVir-D2178-1395/2020|EPI_IS ···················· ···················· ···················· ···················· ······T············· ····················
AA 6479 hCoV-19/Germany/DE-BE-ChVir-D749-5790/2020|EPI_ISL ···················· ···················· ···················· ···················· ···················· ····················
AA 6480 hCoV-19/Germany/DE-BE-ChVir-D762-6625/2020|EPI_ISL ···················· ···················· ···················· ···················· ···················· ····················
AA 6481 hCoV-19/Germany/DE-BE-ChVir-D792-7028/2020|EPI_ISL ···················· ···················· ···················· ···················· ···················· ····················
AA 6482 hCoV-19/Germany/DE-BE-ChVir-D793-8179/2020|EPI_ISL ···················· ···················· ···················· ···················· ···················· ····················
AA 6483 hCoV-19/Germany/DE-BE-ChVir-D903-4341/2020|EPI_ISL ···················· ···················· ···················· ···················· ···················· ····················
AA 6484 hCoV-19/Germany/DE-BE-ChVir-D909-4341/2020|EPI_ISL ···················· ···················· ···················· ···················· ···················· ····················
AA 6485 hCoV-19/Germany/DE-BE-ChVir-W1042-8106/2020|EPI_IS ···················· ···················· ···················· ···················· ···················· ····················
AA 6486 hCoV-19/Germany/DE-BE-ChVir-W1043-8183/2020|EPI_IS ···················· ···················· ···················· ···················· ···················· ····················
AA 6487 hCoV-19/Germany/DE-BE-ChVir-W1045-5646/2020|EPI_IS ···················· ···················· ···················· ···················· ···················· ····················
AA 6488 hCoV-19/Germany/DE-BE-ChVir-W1048-4346/2020|EPI_IS ···················· ···················· ···················· ···················· ···················· ····················
AA 6489 hCoV-19/Germany/DE-BE-ChVir-W1055-8157/2020|EPI_IS ···················· ···················· ···················· ···················· ···················· ····················
AA 6490 hCoV-19/Germany/DE-BE-ChVir-W1060-4393/2020|EPI_IS ···················· ···················· ···················· ···················· ···················· ····················
AA 6491 hCoV-19/Germany/DE-BE-ChVir-W1064-4324/2020|EPI_IS ···················· ···················· ···················· ···················· ···················· ····················
AA 6492 hCoV-19/Germany/DE-BE-ChVir-W1072-4344/2020|EPI_IS ···················· ···················· ···················· ···················· ···················· ····················
AA 6493 hCoV-19/Germany/DE-BE-ChVir-W1445-6284/2020|EPI_IS ···················· ···················· ···················· ···················· ···················· ····················
AA 6494 hCoV-19/USA/NY-Wadsworth-263697-01/2020|EPI_ISL_75 ···················· ···················· ···················· ···················· ···················· ····················

| | | | | | | | | |
|---|---|---|---|---|---|---|---|---|
| AA | 6495 | hCoV-19/USA/NY-Wadsworth-267180-01/2020\|EPI_ISL_75 | ···················· | ···················· | ····················· | ···················· | ···················· | ···················· |
| AA | 6496 | hCoV-19/USA/NY-Wadsworth-280404-01/2020\|EPI_ISL_75 | ···················· | ·A·················· | ····················· | ···················· | ···················· | ···················· |
| AA | 6497 | hCoV-19/USA/NY-Wadsworth-282089-01/2020\|EPI_ISL_75 | ···················· | ···················· | ····················· | ···················· | ···················· | ···················· |
| AA | 6498 | hCoV-19/USA/NY-Wadsworth-283214-01/2020\|EPI_ISL_75 | ···················· | ···················· | ····················· | ···················· | ···················· | ···················· |
| AA | 6499 | hCoV-19/USA/NY-Wadsworth-281848-01/2020\|EPI_ISL_75 | ···················· | ···················· | ····················· | ···················· | ···················· | ···················· |
| AA | 6500 | hCoV-19/USA/NY-Wadsworth-281309-01/2020\|EPI_ISL_75 | ···················· | ···················· | ····················· | ···················· | ···················· | ···················· |
| AA | 6501 | hCoV-19/USA/NY-Wadsworth-280623-01/2020\|EPI_ISL_75 | ···················· | ···················· | ····················· | ···················· | ···················· | ···················· |
| AA | 6502 | hCoV-19/USA/NY-Wadsworth-280624-01/2020\|EPI_ISL_75 | ···················· | ···················· | ····················· | ···················· | ···················· | ···················· |
| AA | 6503 | hCoV-19/USA/NY-Wadsworth-281613-01/2020\|EPI_ISL_75 | ···················· | ···················· | ····················· | ···················· | ···················· | ···················· |
| AA | 6504 | hCoV-19/USA/NY-Wadsworth-281938-01/2020\|EPI_ISL_75 | ···················· | ·A·················· | ····················· | ···················· | ···················· | ···················· |
| AA | 6505 | hCoV-19/USA/NY-Wadsworth-281937-01/2020\|EPI_ISL_75 | ···················· | ·A·················· | ····················· | ···················· | ···················· | ···················· |
| AA | 6506 | hCoV-19/USA/NY-Wadsworth-281984-01/2020\|EPI_ISL_75 | ···················· | ·A·················· | ····················· | ···················· | ···················· | ···················· |
| AA | 6507 | hCoV-19/USA/NY-Wadsworth-281986-01/2020\|EPI_ISL_75 | ···················· | ·A·················· | ····················· | ···················· | ···················· | ···················· |
| AA | 6508 | hCoV-19/USA/NY-Wadsworth-282014-01/2020\|EPI_ISL_75 | ···················· | ·A·················· | ····················· | ···················· | ···················· | ···················· |
| AA | 6509 | hCoV-19/USA/NY-Wadsworth-281658-01/2020\|EPI_ISL_75 | ···················· | ···················· | ····················· | ···················· | ···················· | ···················· |
| AA | 6510 | hCoV-19/USA/NY-Wadsworth-281619-01/2020\|EPI_ISL_75 | ···················· | ·A·················· | ····················· | ···················· | ···················· | ···················· |
| AA | 6511 | hCoV-19/USA/NY-Wadsworth-281624-01/2020\|EPI_ISL_75 | ···················· | ···················· | ····················· | ···················· | ···················· | ···················· |
| AA | 6512 | hCoV-19/USA/NY-Wadsworth-281633-01/2020\|EPI_ISL_75 | ···················· | ·A·················· | ····················· | ···················· | ···················· | ···················· |
| AA | 6513 | hCoV-19/USA/NY-Wadsworth-281667-01/2020\|EPI_ISL_75 | ···················· | ···················· | ····················· | ···················· | ···················· | ···················· |
| AA | 6514 | hCoV-19/USA/NY-Wadsworth-281895-01/2020\|EPI_ISL_75 | ···················· | ···················· | ····················· | ···················· | T··················· | ···················· |
| AA | 6515 | hCoV-19/USA/NY-Wadsworth-281900-01/2020\|EPI_ISL_75 | ···················· | ···················· | ····················· | ···················· | T··················· | ···················· |
| AA | 6516 | hCoV-19/USA/NY-Wadsworth-281870-01/2020\|EPI_ISL_75 | ···················· | ···················· | ····················· | ···················· | T··················· | ···················· |
| AA | 6517 | hCoV-19/USA/NY-Wadsworth-281871-01/2020\|EPI_ISL_75 | ···················· | ···················· | ····················· | ···················· | T··················· | ···················· |
| AA | 6518 | hCoV-19/USA/NY-Wadsworth-281872-01/2020\|EPI_ISL_75 | ···················· | ···················· | ····················· | ···················· | T··················· | ···················· |
| AA | 6519 | hCoV-19/USA/NY-Wadsworth-281873-01/2020\|EPI_ISL_75 | ···················· | ···················· | ····················· | ···················· | T··················· | ···················· |
| AA | 6520 | hCoV-19/USA/NY-Wadsworth-282098-01/2020\|EPI_ISL_75 | ···················· | ···················· | ····················· | ···················· | ···················· | ···················· |
| AA | 6521 | hCoV-19/USA/NY-Wadsworth-282596-01/2020\|EPI_ISL_75 | ···················· | ·A·················· | ····················· | ···················· | ···················· | ···················· |
| AA | 6522 | hCoV-19/USA/NY-Wadsworth-282646-01/2020\|EPI_ISL_75 | ···················· | ···················· | ····················· | ···················· | ···················· | ···················· |
| AA | 6523 | hCoV-19/USA/NY-Wadsworth-282647-01/2020\|EPI_ISL_75 | ···················· | ···················· | ····················· | ···················· | ···················· | ···················· |
| AA | 6524 | hCoV-19/USA/NY-Wadsworth-282648-01/2020\|EPI_ISL_75 | ···················· | ···················· | ····················· | ···················· | ···················· | ···················· |
| AA | 6525 | hCoV-19/USA/NY-Wadsworth-282631-01/2020\|EPI_ISL_75 | ···················· | ···················· | ····················· | ···················· | ···················· | ···················· |
| AA | 6526 | hCoV-19/USA/NY-Wadsworth-282632-01/2020\|EPI_ISL_75 | ···················· | ···················· | ····················· | ···················· | ···················· | ···················· |
| AA | 6527 | hCoV-19/USA/NY-Wadsworth-282645-01/2020\|EPI_ISL_75 | ···················· | ···················· | ····················· | ···················· | ···················· | ···················· |
| AA | 6528 | hCoV-19/USA/NY-Wadsworth-282305-01/2020\|EPI_ISL_75 | ···················· | ···················· | ····················· | ···················· | ···················· | ···················· |
| AA | 6529 | hCoV-19/USA/NY-Wadsworth-285049-01/2020\|EPI_ISL_75 | ···················· | ···················· | ····················· | ···················· | ···················· | ···················· |
| AA | 6530 | hCoV-19/USA/NY-Wadsworth-284996-01/2020\|EPI_ISL_75 | ···················· | ···················· | ····················· | ···················· | ···················· | ···················· |
| AA | 6531 | hCoV-19/USA/NY-Wadsworth-284999-01/2020\|EPI_ISL_75 | ···················· | ···················· | ····················· | ···················· | ···················· | ···················· |
| AA | 6532 | hCoV-19/USA/NY-Wadsworth-285036-01/2020\|EPI_ISL_75 | ···················· | ···················· | ····················· | ···················· | ···················· | ···················· |
| AA | 6533 | hCoV-19/USA/NY-Wadsworth-285337-01/2020\|EPI_ISL_75 | ···················· | ·A·················· | ····················· | ···················· | ···················· | ···················· |
| AA | 6534 | hCoV-19/USA/NY-Wadsworth-285291-01/2020\|EPI_ISL_75 | ···················· | ···················· | ············G········ | ···················· | ···················· | ···················· |
| AA | 6535 | hCoV-19/USA/NY-Wadsworth-285388-01/2020\|EPI_ISL_75 | ···················· | ···················· | ····················· | ···················· | ···················· | ···················· |
| AA | 6536 | hCoV-19/USA/NY-Wadsworth-278554-01/2020\|EPI_ISL_75 | ···················· | ···················· | ····················· | ···················· | ···················· | ···················· |
| AA | 6537 | hCoV-19/USA/NY-Wadsworth-279166-01/2020\|EPI_ISL_75 | ···················· | ···················· | ····················· | ···················· | ···················· | ···················· |
| AA | 6538 | hCoV-19/USA/NY-Wadsworth-279142-01/2020\|EPI_ISL_75 | ···················· | ···················· | ····················· | ···················· | ···················· | ···················· |
| AA | 6539 | hCoV-19/USA/NY-Wadsworth-279198-01/2020\|EPI_ISL_75 | ···················· | ···················· | ····················· | ···················· | ···················· | ···················· |
| AA | 6540 | hCoV-19/USA/NY-Wadsworth-278894-01/2020\|EPI_ISL_75 | ···················· | ·A·················· | ····················· | ···················· | ···················· | ···················· |
| AA | 6541 | hCoV-19/USA/NY-Wadsworth-279611-01/2020\|EPI_ISL_75 | ···················· | ·A·················· | ····················· | ···················· | ···················· | ···················· |
| AA | 6542 | hCoV-19/USA/NY-Wadsworth-279500-01/2020\|EPI_ISL_75 | ···················· | ·A·················· | ····················· | ···················· | ···················· | ···················· |
| AA | 6543 | hCoV-19/USA/NY-Wadsworth-279778-01/2020\|EPI_ISL_75 | ···················· | ···················· | ····················· | ···················· | ···················· | ···················· |
| AA | 6544 | hCoV-19/USA/NY-Wadsworth-279841-01/2020\|EPI_ISL_75 | ···················· | ···················· | ····················· | ···················· | ···················· | ···················· |
| AA | 6545 | hCoV-19/USA/NY-Wadsworth-279952-01/2020\|EPI_ISL_75 | ···················· | ···················· | ····················· | ···················· | ···················· | ···················· |
| AA | 6546 | hCoV-19/USA/NY-Wadsworth-279587-01/2020\|EPI_ISL_75 | ···················· | ·A·················· | ····················· | ···················· | ···················· | ···················· |
| AA | 6547 | hCoV-19/USA/NY-Wadsworth-280011-01/2020\|EPI_ISL_75 | ···················· | ·A·················· | ····················· | ···················· | ···················· | ···················· |
| AA | 6548 | hCoV-19/USA/NY-Wadsworth-280303-01/2020\|EPI_ISL_75 | ···················· | ···················· | ····················· | ···················· | ···················· | ···················· |
| AA | 6549 | hCoV-19/USA/NY-Wadsworth-280302-01/2020\|EPI_ISL_75 | ···················· | ···················· | ····················· | ···················· | ···················· | ···················· |
| AA | 6550 | hCoV-19/USA/NY-Wadsworth-280082-01/2020\|EPI_ISL_75 | ···················· | ·A·················· | ····················· | ···················· | ···················· | ···················· |
| AA | 6551 | hCoV-19/USA/NY-Wadsworth-280102-01/2020\|EPI_ISL_75 | ···················· | ···················· | ····················· | ···················· | ···················· | ···················· |
| AA | 6552 | hCoV-19/USA/NY-Wadsworth-280264-01/2020\|EPI_ISL_75 | ···················· | ···················· | ····················· | ···················· | ···················· | ···················· |
| AA | 6553 | hCoV-19/USA/NY-Wadsworth-280101-01/2020\|EPI_ISL_75 | ···················· | ···················· | ····················· | ···················· | ···················· | ···················· |
| AA | 6554 | hCoV-19/USA/NY-Wadsworth-280041-01/2020\|EPI_ISL_75 | ···················· | ···················· | ····················· | ···················· | ···················· | ···················· |
| AA | 6555 | hCoV-19/USA/NY-Wadsworth-280025-01/2020\|EPI_ISL_75 | ···················· | ···················· | ····················· | ···················· | ···················· | ···················· |
| AA | 6556 | hCoV-19/USA/NY-Wadsworth-280026-01/2020\|EPI_ISL_75 | ···················· | ···················· | ····················· | ···················· | ···················· | ···················· |
| AA | 6557 | hCoV-19/USA/NY-Wadsworth-280024-01/2020\|EPI_ISL_75 | ···················· | ···················· | ····················· | ···················· | ···················· | ···················· |
| AA | 6558 | hCoV-19/USA/NY-Wadsworth-280499-01/2020\|EPI_ISL_75 | ···················· | ···················· | ····················· | ···················· | ···················· | ···················· |
| AA | 6559 | hCoV-19/Canada/ON-S1987/2020\|EPI_ISL_755700\|2020-1 | ···················· | ···················· | ····················· | ···················· | ···················· | ···················· |
| AA | 6560 | hCoV-19/Canada/ON-S1988/2020\|EPI_ISL_755701\|2020-1 | ···················· | ·A·················· | ····················· | ···················· | ···················· | ···A················ |
| AA | 6561 | hCoV-19/Canada/ON-S1991/2020\|EPI_ISL_755702\|2020-1 | ···················· | ·A·················· | ····················· | ···················· | ···················· | ···A················ |
| AA | 6562 | hCoV-19/Canada/ON-S1992/2020\|EPI_ISL_755703\|2020-1 | ···················· | ···················· | ····················· | ···················· | ···················· | ···················· |
| AA | 6563 | hCoV-19/Canada/ON-S1995/2020\|EPI_ISL_755705\|2020-1 | ···················· | ·A·················· | ····················· | ···················· | ···················· | ···A················ |
| AA | 6564 | hCoV-19/Canada/ON-S1996/2020\|EPI_ISL_755706\|2020-1 | ···················· | ···················· | ····················· | ···················· | ···················· | ···················· |
| AA | 6565 | hCoV-19/Canada/ON-S1997/2020\|EPI_ISL_755707\|2020-1 | ···················· | ···················· | ····················· | ···················· | ···················· | ···················· |
| AA | 6566 | hCoV-19/Canada/ON-S2004/2020\|EPI_ISL_755709\|2020-1 | ···················· | ···················· | ····················· | ···················· | ···················· | ···················· |
| AA | 6567 | hCoV-19/Canada/ON-S2005/2020\|EPI_ISL_755710\|2020-1 | ···················· | ···················· | ····················· | ···················· | ···················· | ···················· |
| AA | 6568 | hCoV-19/Canada/ON-S2006/2020\|EPI_ISL_755711\|2020-1 | ···················· | ···················· | ····················· | ···················· | ···················· | ···················· |
| AA | 6569 | hCoV-19/Canada/ON-A1/2020\|EPI_ISL_755712\|2020-11-2 | ···················· | ···················· | ····················· | ···················· | ···················· | ···················· |
| AA | 6570 | hCoV-19/Canada/ON-A10/2020\|EPI_ISL_755713\|2020-11- | ···················· | ·A·················· | ····················· | ···················· | ···················· | ···A················ |
| AA | 6571 | hCoV-19/Canada/ON-A11/2020\|EPI_ISL_755714\|2020-11- | ···················· | ·A·················· | ····················· | ···················· | ···················· | ···A················ |
| AA | 6572 | hCoV-19/Canada/ON-A12/2020\|EPI_ISL_755715\|2020-11- | ···················· | ·A·················· | ····················· | ···················· | ···················· | ···A················ |
| AA | 6573 | hCoV-19/Canada/ON-A13/2020\|EPI_ISL_755716\|2020-11- | ···················· | ···················· | ····················· | ···················· | ···················· | ···················· |
| AA | 6574 | hCoV-19/Canada/ON-A15/2020\|EPI_ISL_755717\|2020-11- | ···················· | ·A·················· | ····················· | ···················· | ···················· | ···A················ |

| | | | | | | | | |
|---|---|---|---|---|---|---|---|---|
| AA | 6575 | hCoV-19/Canada/ON-A16/2020\|EPI_ISL_755718\|2020-11- | ···················· | ···················· | ····················· | ···················· | ···················· | ···················· |
| AA | 6576 | hCoV-19/Canada/ON-A19/2020\|EPI_ISL_755719\|2020-11- | ···················· | ·A·················· | ····················· | ···················· | ···················· | ···A················ |
| AA | 6577 | hCoV-19/Canada/ON-A2/2020\|EPI_ISL_755720\|2020-11-2 | ···················· | ·A·················· | ····················· | ···················· | ···················· | ···A················ |
| AA | 6578 | hCoV-19/Canada/ON-A21/2020\|EPI_ISL_755721\|2020-11- | ···················· | ···················· | ····················· | ···················· | ···················· | ···················· |
| AA | 6579 | hCoV-19/Canada/ON-A23/2020\|EPI_ISL_755722\|2020-11- | ···················· | ·A·················· | ····················· | ···················· | ···················· | ···A················ |
| AA | 6580 | hCoV-19/Canada/ON-A25/2020\|EPI_ISL_755723\|2020-11- | ···················· | ·A·················· | ····················· | ···················· | ···················· | ···A················ |
| AA | 6581 | hCoV-19/Canada/ON-A26/2020\|EPI_ISL_755724\|2020-11- | ···················· | ·A·················· | ····················· | ···················· | ···················· | ···A················ |
| AA | 6582 | hCoV-19/Canada/ON-A27/2020\|EPI_ISL_755725\|2020-11- | ···················· | ·A·················· | ····················· | ···················· | ···················· | ···A················ |
| AA | 6583 | hCoV-19/Canada/ON-A28/2020\|EPI_ISL_755726\|2020-11- | ···················· | ···················· | ····················· | ···················· | ···················· | ···················· |
| AA | 6584 | hCoV-19/Canada/ON-A3/2020\|EPI_ISL_755727\|2020-11-2 | ···················· | ·A·················· | ····················· | ···················· | ···················· | ···A················ |
| AA | 6585 | hCoV-19/Canada/ON-A30/2020\|EPI_ISL_755728\|2020-11- | ···················· | ·A·················· | ····················· | ···················· | ···················· | ···A················ |
| AA | 6586 | hCoV-19/Canada/ON-A31/2020\|EPI_ISL_755729\|2020-11- | ···················· | ···················· | ····················· | ···················· | ···················· | ···················· |
| AA | 6587 | hCoV-19/Canada/ON-A32/2020\|EPI_ISL_755730\|2020-11- | ···················· | ·A·················· | ····················· | ···················· | ···················· | ···A················ |
| AA | 6588 | hCoV-19/Canada/ON-A4/2020\|EPI_ISL_755731\|2020-11-2 | ···················· | ···················· | ····················· | ···················· | ···················· | ···················· |
| AA | 6589 | hCoV-19/Canada/ON-A6/2020\|EPI_ISL_755733\|2020-11-2 | ···················· | ···················· | ····················· | ···················· | ···················· | ···················· |
| AA | 6590 | hCoV-19/Canada/ON-A7/2020\|EPI_ISL_755734\|2020-11-2 | ···················· | ·A·················· | ····················· | ···················· | ···················· | ···A················ |
| AA | 6591 | hCoV-19/Canada/ON-A8/2020\|EPI_ISL_755735\|2020-11-2 | ···················· | ···················· | ····················· | ···················· | ···················· | ···················· |
| AA | 6592 | hCoV-19/Canada/ON-A9/2020\|EPI_ISL_755736\|2020-11-2 | ···················· | ·A·················· | ····················· | ···················· | ···················· | ···A················ |
| AA | 6593 | hCoV-19/Canada/ON-SB-01/2020\|EPI_ISL_755742\|2020-1 | ···················· | ·A·················· | ····················· | ···················· | ···················· | ···A················ |
| AA | 6594 | hCoV-19/Canada/ON-SB-04/2020\|EPI_ISL_755744\|2020-1 | ···················· | ·A·················· | ····················· | ···················· | ···················· | ···A················ |
| AA | 6595 | hCoV-19/Canada/ON-SB-06/2020\|EPI_ISL_755745\|2020-1 | ···················· | ·A·················· | ····················· | ···················· | ···················· | ···A················ |
| AA | 6596 | hCoV-19/Canada/ON-SB-09/2020\|EPI_ISL_755748\|2020-1 | ···················· | ·A·················· | ····················· | ···················· | ···················· | ···A················ |
| AA | 6597 | hCoV-19/Canada/ON-SB-10/2020\|EPI_ISL_755749\|2020-1 | ···················· | ·A·················· | ····················· | ···················· | ···················· | ···A················ |
| AA | 6598 | hCoV-19/Canada/ON-S1628/2020\|EPI_ISL_755755\|2020-0 | ···················· | ···················· | ····················· | ···················· | ···················· | ···················· |
| AA | 6599 | hCoV-19/Canada/ON-S1629/2020\|EPI_ISL_755756\|2020-0 | ···················· | ···················· | ····················· | ···················· | ···················· | ···················· |
| AA | 6600 | hCoV-19/Canada/ON-S1636/2020\|EPI_ISL_755757\|2020-0 | ···················· | ·A·················· | ····················· | ···················· | ···················· | ···A················ |
| AA | 6601 | hCoV-19/Canada/ON-S1637/2020\|EPI_ISL_755758\|2020-0 | ···················· | ···················· | ····················· | ···················· | ···················· | ···················· |
| AA | 6602 | hCoV-19/Canada/ON-S1760/2020\|EPI_ISL_755761\|2020-0 | ···················· | ·A·················· | ····················· | ···················· | ···················· | ···A················ |
| AA | 6603 | hCoV-19/Canada/ON-S1761/2020\|EPI_ISL_755762\|2020-0 | ···················· | ·A·················· | ····················· | ···················· | ···················· | ···A················ |
| AA | 6604 | hCoV-19/Canada/ON-S1762/2020\|EPI_ISL_755763\|2020-0 | ···················· | ···················· | ····················· | ···················· | ···················· | ···················· |
| AA | 6605 | hCoV-19/Canada/ON-S1763/2020\|EPI_ISL_755764\|2020-0 | ···················· | ·A·················· | ····················· | ···················· | ···················· | ···A················ |
| AA | 6606 | hCoV-19/Canada/ON-S1764/2020\|EPI_ISL_755765\|2020-0 | ···················· | ···················· | ····················· | ···················· | ···················· | ···················· |
| AA | 6607 | hCoV-19/Canada/ON-S1765/2020\|EPI_ISL_755766\|2020-0 | ···················· | ···················· | ····················· | ···················· | ···················· | ···················· |
| AA | 6608 | hCoV-19/Finland/15LS14AF1/2020\|EPI_ISL_756170\|2020 | ···················· | ···················· | ····················· | ···················· | ···················· | ···················· |
| AA | 6609 | hCoV-19/Finland/15LS3AA11/2020\|EPI_ISL_756171\|2020 | ···················· | ·A·················· | ····················· | ···················· | ···················· | ···A················ |
| AA | 6610 | hCoV-19/Finland/15LS3AA2/2020\|EPI_ISL_756172\|2020- | ···················· | ···················· | ····················· | ···················· | ···················· | ···················· |
| AA | 6611 | hCoV-19/Finland/15LS3AE3/2020\|EPI_ISL_756173\|2020- | ···················· | ···················· | ····················· | ···················· | ···················· | ···················· |
| AA | 6612 | hCoV-19/Finland/15LS4BD7/2020\|EPI_ISL_756174\|2020- | ···················· | ···················· | ····················· | ···················· | ···················· | ···················· |
| AA | 6613 | hCoV-19/Finland/15LS4BF5/2020\|EPI_ISL_756175\|2020- | ···················· | ·A·················· | ····················· | ···················· | ···················· | ···················· |
| AA | 6614 | hCoV-19/Finland/15LS4BG11/2020\|EPI_ISL_756176\|2020 | ···················· | ·A·················· | ····················· | ···················· | ···················· | ···A················ |
| AA | 6615 | hCoV-19/Finland/15LS6BB7/2020\|EPI_ISL_756177\|2020- | ···················· | ·A·················· | ····················· | ···················· | ···················· | ···A················ |
| AA | 6616 | hCoV-19/Finland/15LS6BE3/2020\|EPI_ISL_756178\|2020- | ···················· | ·A·················· | ····················· | ···················· | ···················· | ···A················ |
| AA | 6617 | hCoV-19/Finland/15LS7AA11/2020\|EPI_ISL_756179\|2020 | ···················· | ···················· | ····················· | ···················· | ···················· | ···················· |
| AA | 6618 | hCoV-19/Finland/15LS7AF9/2020\|EPI_ISL_756180\|2020- | ···················· | ···················· | ····················· | ···················· | ···················· | ···················· |
| AA | 6619 | hCoV-19/Finland/15SS14H3/2020\|EPI_ISL_756181\|2020- | ···················· | ···················· | ····················· | ···················· | ···················· | ···················· |
| AA | 6620 | hCoV-19/Finland/15SS20B12/2020\|EPI_ISL_756182\|2020 | ···················· | ···················· | ····················· | ···················· | ···················· | ···················· |
| AA | 6621 | hCoV-19/Finland/15SS9H3/2020\|EPI_ISL_756183\|2020-0 | ···················· | ···················· | ····················· | ···················· | ···················· | ···················· |
| AA | 6622 | hCoV-19/USA/WY-WPHL-20157383/2020\|EPI_ISL_756184\|2 | ···················· | ···················· | ····················· | ···················· | ···················· | ···················· |
| AA | 6623 | hCoV-19/USA/WY-WPHL-20157466/2020\|EPI_ISL_756185\|2 | ···················· | ···················· | ····················· | ···················· | ···················· | ···················· |
| AA | 6624 | hCoV-19/USA/WY-WPHL-20157548/2020\|EPI_ISL_756186\|2 | ···················· | ·A·················· | ····················· | ···················· | ···················· | ···················· |
| AA | 6625 | hCoV-19/USA/WY-WPHL-20157593/2020\|EPI_ISL_756187\|2 | ···················· | ···················· | ····················· | ···················· | ···················· | ···················· |
| AA | 6626 | hCoV-19/USA/WY-WPHL-20157594/2020\|EPI_ISL_756188\|2 | ···················· | ···················· | ····················· | ···················· | ···················· | ···················· |
| AA | 6627 | hCoV-19/USA/WY-WPHL-20157618/2020\|EPI_ISL_756189\|2 | ···················· | ···················· | ····················· | ···················· | ···················· | ···················· |
| AA | 6628 | hCoV-19/USA/WY-WPHL-20157685/2020\|EPI_ISL_756190\|2 | ···················· | ·A·················· | ····················· | ···················· | ···················· | ···················· |
| AA | 6629 | hCoV-19/USA/WA-UW-46833/2020\|EPI_ISL_756191\|2020-1 | ···················· | ···················· | ····················· | ···················· | ···················· | ···················· |
| AA | 6630 | hCoV-19/USA/WA-UW-46835/2020\|EPI_ISL_756192\|2020-1 | ···················· | ···················· | ····················· | ···················· | ···················· | ···················· |
| AA | 6631 | hCoV-19/USA/WA-UW-46839/2020\|EPI_ISL_756194\|2020-1 | ···················· | ·A·················· | ····················· | ···················· | ···················· | ···················· |
| AA | 6632 | hCoV-19/USA/WA-UW-46863/2020\|EPI_ISL_756195\|2020-1 | ···················· | ···················· | ····················· | ···················· | ···················· | ···················· |
| AA | 6633 | hCoV-19/USA/WA-UW-46868/2020\|EPI_ISL_756196\|2020-1 | ········T··········· | ·A·················· | ····················· | ···················· | ···················· | ···················· |
| AA | 6634 | hCoV-19/USA/WA-UW-46871/2020\|EPI_ISL_756197\|2020-1 | ···················· | ···················· | ····················· | ···················· | ···················· | ···················· |
| AA | 6635 | hCoV-19/USA/WA-UW-46880/2020\|EPI_ISL_756198\|2020-1 | ···················· | ···················· | ····················· | ···················· | ···················· | ···················· |
| AA | 6636 | hCoV-19/USA/WA-UW-46885/2020\|EPI_ISL_756199\|2020-1 | ···················· | ···················· | ····················· | ···················· | ···················· | ···················· |
| AA | 6637 | hCoV-19/Denmark/DCGC-20657/2020\|EPI_ISL_759280\|202 | ···················· | ···················· | ····················· | ···················· | ···················· | ···················· |
| AA | 6638 | hCoV-19/Denmark/DCGC-20658/2020\|EPI_ISL_759281\|202 | ···················· | ···················· | ····················· | ···················· | ···················· | ···················· |
| AA | 6639 | hCoV-19/Denmark/DCGC-20659/2020\|EPI_ISL_759282\|202 | ···················· | ···················· | ····················· | ···················· | ···················· | ···················· |
| AA | 6640 | hCoV-19/Denmark/DCGC-20661/2020\|EPI_ISL_759283\|202 | ···················· | ···················· | ····················· | ···················· | ···················· | ···················· |
| AA | 6641 | hCoV-19/Denmark/DCGC-20663/2020\|EPI_ISL_759284\|202 | ···················· | ···················· | ····················· | ···················· | ···················· | ···················· |
| AA | 6642 | hCoV-19/Denmark/DCGC-20665/2020\|EPI_ISL_759285\|202 | ···················· | ···················· | ····················· | ···················· | ···C················ | ···················· |
| AA | 6643 | hCoV-19/Denmark/DCGC-20668/2020\|EPI_ISL_759286\|202 | ···················· | ···················· | ····················· | ···················· | ···················· | ···················· |
| AA | 6644 | hCoV-19/Denmark/DCGC-20669/2020\|EPI_ISL_759287\|202 | ···················· | ···················· | ····················· | ···················· | ···················· | ···················· |
| AA | 6645 | hCoV-19/Denmark/DCGC-20670/2020\|EPI_ISL_759288\|202 | ···················· | ···················· | ····················· | ···················· | ···················· | ···················· |
| AA | 6646 | hCoV-19/Denmark/DCGC-20672/2020\|EPI_ISL_759289\|202 | ···················· | ···················· | ····················· | ···················· | ···················· | ···················· |
| AA | 6647 | hCoV-19/Denmark/DCGC-20673/2020\|EPI_ISL_759290\|202 | ···················· | ···················· | ····················· | ···················· | ···················· | ···················· |
| AA | 6648 | hCoV-19/Denmark/DCGC-20674/2020\|EPI_ISL_759291\|202 | ···················· | ···················· | ····················· | ···················· | ···················· | ···················· |
| AA | 6649 | hCoV-19/Denmark/DCGC-20675/2020\|EPI_ISL_759292\|202 | ···················· | ···················· | ····················· | ···················· | ···················· | ···················· |
| AA | 6650 | hCoV-19/Denmark/DCGC-20676/2020\|EPI_ISL_759293\|202 | ···················· | ···················· | ····················· | ···················· | ···················· | ···················· |
| AA | 6651 | hCoV-19/Denmark/DCGC-20677/2020\|EPI_ISL_759294\|202 | ···················· | ···················· | ····················· | ···················· | ···················· | ···················· |
| AA | 6652 | hCoV-19/Denmark/DCGC-20679/2020\|EPI_ISL_759295\|202 | ···················· | ···················· | ····················· | ···················· | ···················· | ···················· |
| AA | 6653 | hCoV-19/Denmark/DCGC-20681/2020\|EPI_ISL_759296\|202 | ···················· | ···················· | ····················· | ···················· | ···················· | ···················· |
| AA | 6654 | hCoV-19/Denmark/DCGC-20682/2020\|EPI_ISL_759297\|202 | ···················· | ···················· | ····················· | ···················· | ···················· | ···················· |

| | | | | | | | |
|---|---|---|---|---|---|---|---|
| AA | 6655 hCoV-19/Denmark/DCGC-20683/2020\|EPI_ISL_759298\|202 | ···················· | ···················· | ···················· | ···················· | ···················· | ···················· |
| AA | 6656 hCoV-19/Denmark/DCGC-20685/2020\|EPI_ISL_759299\|202 | ···················· | ···················· | ···················· | ···················· | ···················· | ···················· |
| AA | 6657 hCoV-19/South Korea/KDCA0092/2020\|EPI_ISL_747331\|2 | ···················· | ···················· | ···················· | ···················· | ···················· | ···················· |
| AA | 6658 hCoV-19/South Korea/KDCA0094/2020\|EPI_ISL_747332\|2 | ···················· | ···················· | ···················· | ···················· | ···················· | ···················· |
| AA | 6659 hCoV-19/South Korea/KDCA0100/2020\|EPI_ISL_747336\|2 | ···················· | ···················· | ···················· | ···················· | ···················· | ···················· |
| AA | 6660 hCoV-19/South Korea/KDCA0101/2020\|EPI_ISL_747337\|2 | ···················· | ···················· | ···················· | ···················· | ···················· | ···················· |
| AA | 6661 hCoV-19/South Korea/KDCA0102/2020\|EPI_ISL_747338\|2 | ···················· | ···················· | ···················· | ···················· | ···················· | ···················· |
| AA | 6662 hCoV-19/South Korea/KDCA0103/2020\|EPI_ISL_747339\|2 | ···················· | ···················· | ···················· | ···················· | ···················· | ···················· |
| AA | 6663 hCoV-19/Denmark/DCGC-18857/2020\|EPI_ISL_748600\|202 | ···················· | ···················· | ···················· | ···················· | ···················· | ···················· |
| AA | 6664 hCoV-19/Denmark/DCGC-18859/2020\|EPI_ISL_748601\|202 | ···················· | ···················· | ···················· | ···················· | ···················· | ···················· |
| AA | 6665 hCoV-19/Denmark/DCGC-18860/2020\|EPI_ISL_748602\|202 | ···················· | ···················· | ···················· | ···················· | ···················· | ···················· |
| AA | 6666 hCoV-19/Denmark/DCGC-18861/2020\|EPI_ISL_748603\|202 | ···················· | ···················· | ···················· | ···················· | ···················· | ···················· |
| AA | 6667 hCoV-19/Denmark/DCGC-18863/2020\|EPI_ISL_748604\|202 | ···················· | ···················· | ···················· | ···················· | ···················· | ···················· |
| AA | 6668 hCoV-19/Denmark/DCGC-18864/2020\|EPI_ISL_748605\|202 | ···················· | ·A·················· | ···················· | ···················· | ···················· | ··A················· |
| AA | 6669 hCoV-19/Denmark/DCGC-18865/2020\|EPI_ISL_748606\|202 | ···················· | ···················· | ···················· | ···················· | ···················· | ···················· |
| AA | 6670 hCoV-19/Denmark/DCGC-18867/2020\|EPI_ISL_748607\|202 | ···················· | ···················· | ···················· | ···················· | ···················· | ···················· |
| AA | 6671 hCoV-19/Denmark/DCGC-18871/2020\|EPI_ISL_748608\|202 | ···················· | ···················· | ···················· | ···················· | ···················· | ···················· |
| AA | 6672 hCoV-19/Denmark/DCGC-18873/2020\|EPI_ISL_748609\|202 | ···················· | ···················· | ···················· | ···················· | ···················· | ···················· |
| AA | 6673 hCoV-19/Denmark/DCGC-18875/2020\|EPI_ISL_748610\|202 | ···················· | ···················· | ···················· | ···················· | ···················· | ···················· |
| AA | 6674 hCoV-19/Denmark/DCGC-18876/2020\|EPI_ISL_748611\|202 | ···················· | ···················· | ···················· | ···················· | ···················· | ···················· |
| AA | 6675 hCoV-19/Denmark/DCGC-18877/2020\|EPI_ISL_748612\|202 | ···················· | ···················· | ···················· | ···················· | ···················· | ···················· |
| AA | 6676 hCoV-19/Denmark/DCGC-18878/2020\|EPI_ISL_748613\|202 | ···················· | ···················· | ···················· | ···················· | ···················· | ···················· |
| AA | 6677 hCoV-19/Denmark/DCGC-18880/2020\|EPI_ISL_748614\|202 | ···················· | ···················· | ···················· | ···················· | ···················· | ···················· |
| AA | 6678 hCoV-19/Denmark/DCGC-18881/2020\|EPI_ISL_748615\|202 | ···················· | ···················· | ···················· | ···················· | ···················· | ···················· |
| AA | 6679 hCoV-19/Denmark/DCGC-18884/2020\|EPI_ISL_748616\|202 | ···················· | ···················· | ···················· | ···················· | ···················· | ···················· |
| AA | 6680 hCoV-19/Denmark/DCGC-18885/2020\|EPI_ISL_748617\|202 | ···················· | ···················· | ···················· | ···················· | ···················· | ···················· |
| AA | 6681 hCoV-19/Denmark/DCGC-18886/2020\|EPI_ISL_748618\|202 | ···················· | ···················· | ···················· | ···················· | ···················· | ···················· |
| AA | 6682 hCoV-19/Denmark/DCGC-18887/2020\|EPI_ISL_748619\|202 | ···················· | ···················· | ···················· | ···················· | ···················· | ···················· |
| AA | 6683 hCoV-19/Denmark/DCGC-18889/2020\|EPI_ISL_748620\|202 | ···················· | ···················· | ···················· | ···················· | ···················· | ···················· |
| AA | 6684 hCoV-19/Denmark/DCGC-18890/2020\|EPI_ISL_748621\|202 | ···················· | ···················· | ···················· | ···················· | ···················· | ···················· |
| AA | 6685 hCoV-19/Denmark/DCGC-18892/2020\|EPI_ISL_748622\|202 | ···················· | ···················· | ···················· | ···················· | ···················· | ···················· |
| AA | 6686 hCoV-19/Denmark/DCGC-18893/2020\|EPI_ISL_748623\|202 | ···················· | ···················· | ···················· | ···················· | ···················· | ···················· |
| AA | 6687 hCoV-19/Denmark/DCGC-18895/2020\|EPI_ISL_748624\|202 | ···················· | ···················· | ···················· | ···················· | ···················· | ···················· |
| AA | 6688 hCoV-19/Denmark/DCGC-18896/2020\|EPI_ISL_748625\|202 | ···················· | ···················· | ···················· | ···················· | ···················· | ···················· |
| AA | 6689 hCoV-19/Denmark/DCGC-18898/2020\|EPI_ISL_748626\|202 | ···················· | ···················· | ···················· | ···················· | ···················· | ···················· |
| AA | 6690 hCoV-19/Denmark/DCGC-18900/2020\|EPI_ISL_748627\|202 | ···················· | ···················· | ···················· | ···················· | ···················· | ···················· |
| AA | 6691 hCoV-19/Denmark/DCGC-18901/2020\|EPI_ISL_748628\|202 | ···················· | ···················· | ··A················· | ···················· | ···················· | ···················· |
| AA | 6692 hCoV-19/Denmark/DCGC-18952/2020\|EPI_ISL_748661\|202 | ···················· | ···················· | ···················· | ···················· | ···················· | ···················· |
| AA | 6693 hCoV-19/Denmark/DCGC-18954/2020\|EPI_ISL_748662\|202 | ···················· | ···················· | ···················· | ···················· | ···················· | ···················· |
| AA | 6694 hCoV-19/Denmark/DCGC-18955/2020\|EPI_ISL_748663\|202 | ···················· | ···················· | ···················· | ···················· | ···················· | ···················· |
| AA | 6695 hCoV-19/Denmark/DCGC-18957/2020\|EPI_ISL_748664\|202 | ···················· | ···················· | ···················· | ···················· | ···················· | ···················· |
| AA | 6696 hCoV-19/Denmark/DCGC-18959/2020\|EPI_ISL_748665\|202 | ···················· | ···················· | ···················· | ···················· | ···················· | ···················· |
| AA | 6697 hCoV-19/Denmark/DCGC-18960/2020\|EPI_ISL_748666\|202 | ···················· | ···················· | ···················· | ···················· | ···················· | ···················· |
| AA | 6698 hCoV-19/Uruguay/RIV-M40/2020\|EPI_ISL_748667\|2020-0 | ···················· | ···················· | ···················· | ···················· | ···················· | ···················· |
| AA | 6699 hCoV-19/Denmark/DCGC-18961/2020\|EPI_ISL_748668\|202 | ···················· | ···················· | ···················· | ···················· | ···················· | ···················· |
| AA | 6700 hCoV-19/Denmark/DCGC-18963/2020\|EPI_ISL_748669\|202 | ···················· | ···················· | ···················· | ···················· | ···················· | ···················· |
| AA | 6701 hCoV-19/Denmark/DCGC-18341/2020\|EPI_ISL_749900\|202 | ···················· | ···················· | ···················· | ···················· | ···················· | ···················· |
| AA | 6702 hCoV-19/Denmark/DCGC-18344/2020\|EPI_ISL_749903\|202 | ···················· | ···················· | ···················· | ···················· | ···················· | ···················· |
| AA | 6703 hCoV-19/Denmark/DCGC-18345/2020\|EPI_ISL_749904\|202 | ···················· | ···················· | ···················· | ···················· | ···················· | ···················· |
| AA | 6704 hCoV-19/Denmark/DCGC-18358/2020\|EPI_ISL_749916\|202 | ···················· | ···················· | ···················· | ···················· | ···················· | ···················· |
| AA | 6705 hCoV-19/Denmark/DCGC-18359/2020\|EPI_ISL_749917\|202 | ···················· | ···················· | ···················· | ···················· | ···················· | ···················· |
| AA | 6706 hCoV-19/Denmark/DCGC-18360/2020\|EPI_ISL_749918\|202 | ···················· | ···················· | ···················· | ···················· | ···················· | ···················· |
| AA | 6707 hCoV-19/Denmark/DCGC-18361/2020\|EPI_ISL_749919\|202 | ···················· | ···················· | ···················· | ···················· | ···················· | ···················· |
| AA | 6708 hCoV-19/Denmark/DCGC-18364/2020\|EPI_ISL_749920\|202 | ···················· | ···················· | ···················· | ···················· | ···················· | ···················· |
| AA | 6709 hCoV-19/Denmark/DCGC-18367/2020\|EPI_ISL_749921\|202 | ···················· | ···················· | ···················· | ···················· | ···················· | ···················· |
| AA | 6710 hCoV-19/Denmark/DCGC-18368/2020\|EPI_ISL_749922\|202 | ···················· | ···················· | ···················· | ···················· | ···················· | ···················· |
| AA | 6711 hCoV-19/Denmark/DCGC-18369/2020\|EPI_ISL_749923\|202 | ···················· | ···················· | ···················· | ···················· | ···················· | ···················· |
| AA | 6712 hCoV-19/Denmark/DCGC-18371/2020\|EPI_ISL_749924\|202 | ···················· | ···················· | ···················· | ···················· | ···················· | ···················· |
| AA | 6713 hCoV-19/Denmark/DCGC-18372/2020\|EPI_ISL_749925\|202 | ···················· | ···················· | ···················· | ···················· | ···················· | ···················· |
| AA | 6714 hCoV-19/Denmark/DCGC-18376/2020\|EPI_ISL_749926\|202 | ···················· | ···················· | ···················· | ···················· | ···················· | ···················· |
| AA | 6715 hCoV-19/Denmark/DCGC-18378/2020\|EPI_ISL_749927\|202 | ···················· | ···················· | ···················· | ···················· | ···················· | ···················· |
| AA | 6716 hCoV-19/Denmark/DCGC-18379/2020\|EPI_ISL_749928\|202 | ···················· | ···················· | ···················· | ···················· | ···················· | ···················· |
| AA | 6717 hCoV-19/Denmark/DCGC-18380/2020\|EPI_ISL_749929\|202 | ···················· | ···················· | ···················· | ···················· | ···················· | ···················· |
| AA | 6718 hCoV-19/Denmark/DCGC-18381/2020\|EPI_ISL_749930\|202 | ···················· | ···················· | ···················· | ···················· | ···················· | ···················· |
| AA | 6719 hCoV-19/Denmark/DCGC-18382/2020\|EPI_ISL_749931\|202 | ···················· | ···················· | ···················· | ···················· | ···················· | ···················· |
| AA | 6720 hCoV-19/Denmark/DCGC-18385/2020\|EPI_ISL_749932\|202 | ···················· | ···················· | ···················· | ···················· | ···················· | ···················· |
| AA | 6721 hCoV-19/Denmark/DCGC-18386/2020\|EPI_ISL_749933\|202 | ···················· | ···················· | ···················· | ···················· | ···················· | ···················· |
| AA | 6722 hCoV-19/Denmark/DCGC-18387/2020\|EPI_ISL_749934\|202 | ···················· | ···················· | ···················· | ···················· | ···················· | ···················· |
| AA | 6723 hCoV-19/Denmark/DCGC-18388/2020\|EPI_ISL_749935\|202 | ···················· | ···················· | ···················· | ···················· | ···················· | ···················· |
| AA | 6724 hCoV-19/Denmark/DCGC-18391/2020\|EPI_ISL_749936\|202 | ···················· | ···················· | ···················· | ···················· | ···················· | ···················· |
| AA | 6725 hCoV-19/Denmark/DCGC-18393/2020\|EPI_ISL_749937\|202 | ···················· | ···················· | ···················· | ···················· | ···················· | ···················· |
| AA | 6726 hCoV-19/Denmark/DCGC-18394/2020\|EPI_ISL_749938\|202 | ···················· | ···················· | ···················· | ···················· | ···················· | ···················· |
| AA | 6727 hCoV-19/Denmark/DCGC-18396/2020\|EPI_ISL_749939\|202 | ···················· | ·A·················· | ···················· | ···················· | ···················· | ··A················· |
| AA | 6728 hCoV-19/Denmark/DCGC-18397/2020\|EPI_ISL_749940\|202 | ···················· | ···················· | ···················· | ···················· | ···················· | ···················· |
| AA | 6729 hCoV-19/Denmark/DCGC-18399/2020\|EPI_ISL_749941\|202 | ···················· | ·A·················· | ···················· | ···················· | ···················· | ···················· |
| AA | 6730 hCoV-19/Denmark/DCGC-18400/2020\|EPI_ISL_749942\|202 | ···················· | ···················· | ···················· | ···················· | ···················· | ···················· |
| AA | 6731 hCoV-19/Denmark/DCGC-18402/2020\|EPI_ISL_749943\|202 | ···················· | ···················· | ···················· | ···················· | ···················· | ···················· |
| AA | 6732 hCoV-19/Denmark/DCGC-18403/2020\|EPI_ISL_749944\|202 | ···················· | ·A·················· | ···················· | ···················· | ···················· | ··A················· |
| AA | 6733 hCoV-19/Denmark/DCGC-18404/2020\|EPI_ISL_749945\|202 | ···················· | ···················· | ···················· | ···················· | ···················· | ···················· |
| AA | 6734 hCoV-19/Denmark/DCGC-18406/2020\|EPI_ISL_749946\|202 | ···················· | ···················· | ···················· | ···················· | ···················· | ···················· |

AA 6735 hCoV-19/Denmark/DCGC-18407/2020|EPI_ISL_749947|202 ···················· ···················· ···················· ···················· ···················· ····················
AA 6736 hCoV-19/Denmark/DCGC-18408/2020|EPI_ISL_749948|202 ···················· ···················· ···················· ···················· ··········A········· ····················
AA 6737 hCoV-19/Denmark/DCGC-18409/2020|EPI_ISL_749949|202 ···················· ···················· ···················· ···················· ···················· ····················
AA 6738 hCoV-19/Denmark/DCGC-18410/2020|EPI_ISL_749950|202 ···················· ···················· ···················· ···················· ···················· ····················
AA 6739 hCoV-19/Denmark/DCGC-18412/2020|EPI_ISL_749951|202 ···················· ···················· ···················· ···················· ···················· ····················
AA 6740 hCoV-19/Denmark/DCGC-18413/2020|EPI_ISL_749952|202 ···················· ·A·················· ···················· ···················· ···················· ··A·················
AA 6741 hCoV-19/Denmark/DCGC-18414/2020|EPI_ISL_749953|202 ···················· ···················· ···················· ···················· ···················· ····················
AA 6742 hCoV-19/Denmark/DCGC-18417/2020|EPI_ISL_749954|202 ···················· ···················· ···················· ···················· ···················· ····················
AA 6743 hCoV-19/Denmark/DCGC-18421/2020|EPI_ISL_749955|202 ···················· ···················· ···················· ···················· ···················· ····················
AA 6744 hCoV-19/Denmark/DCGC-18422/2020|EPI_ISL_749956|202 ···················· ···················· ···················· ···················· ···················· ····················
AA 6745 hCoV-19/Denmark/DCGC-18423/2020|EPI_ISL_749957|202 ···················· ···················· ···················· ···················· ···················· ····················
AA 6746 hCoV-19/Denmark/DCGC-18426/2020|EPI_ISL_749958|202 ···················· ···················· ···················· ···················· ···················· ····················
AA 6747 hCoV-19/Denmark/DCGC-18427/2020|EPI_ISL_749959|202 ···················· ·A·················· ···················· ···················· ···················· ··A·················
AA 6748 hCoV-19/Denmark/DCGC-18477/2020|EPI_ISL_749990|202 ···················· ···················· ···················· ···················· ···················· ····················
AA 6749 hCoV-19/Denmark/DCGC-18478/2020|EPI_ISL_749991|202 ···················· ···················· ···················· ···················· ···················· ····················
AA 6750 hCoV-19/Denmark/DCGC-18479/2020|EPI_ISL_749992|202 ···················· ···················· ···················· ···················· ···················· ····················
AA 6751 hCoV-19/Denmark/DCGC-18482/2020|EPI_ISL_749993|202 ···················· ···················· ···················· ···················· ···················· ····················
AA 6752 hCoV-19/Denmark/DCGC-18483/2020|EPI_ISL_749994|202 ···················· ···················· ···················· ···················· ···················· ····················
AA 6753 hCoV-19/Denmark/DCGC-18484/2020|EPI_ISL_749995|202 ···················· ···················· ···················· ···················· ···················· ····················
AA 6754 hCoV-19/Denmark/DCGC-18485/2020|EPI_ISL_749996|202 ···················· ···················· ···················· ···················· ···················· ····················
AA 6755 hCoV-19/Denmark/DCGC-18486/2020|EPI_ISL_749997|202 ···················· ···················· ···················· ···················· ···················· ····················
AA 6756 hCoV-19/Denmark/DCGC-18489/2020|EPI_ISL_749998|202 ···················· ···················· ···················· ···················· ···················· ····················
AA 6757 hCoV-19/Denmark/DCGC-18490/2020|EPI_ISL_749999|202 ···················· ···················· ···················· ···················· ···················· ····················
AA 6758 hCoV-19/Denmark/DCGC-17316/2020|EPI_ISL_750900|202 ···················· ·A·················· ···················· ···················· ···················· ··A·················
AA 6759 hCoV-19/Denmark/DCGC-17317/2020|EPI_ISL_750901|202 ···················· ···················· ···················· ···················· ···················· ····················
AA 6760 hCoV-19/Denmark/DCGC-17318/2020|EPI_ISL_750902|202 ···················· ·A·················· ···················· ···················· ···················· ··A·················
AA 6761 hCoV-19/Denmark/DCGC-17319/2020|EPI_ISL_750903|202 ···················· ···················· ···················· ···················· ···················· ····················
AA 6762 hCoV-19/Denmark/DCGC-17320/2020|EPI_ISL_750904|202 ···················· ···················· ···················· ···················· ···················· ····················
AA 6763 hCoV-19/Denmark/DCGC-17321/2020|EPI_ISL_750905|202 ···················· ·A·················· ···················· ···················· ···················· ··A·················
AA 6764 hCoV-19/Denmark/DCGC-17322/2020|EPI_ISL_750906|202 ···················· ···················· ···················· ···················· ···················· ····················
AA 6765 hCoV-19/Denmark/DCGC-17324/2020|EPI_ISL_750907|202 ···················· ···················· ···················· ···················· ···················· ····················
AA 6766 hCoV-19/Denmark/DCGC-17325/2020|EPI_ISL_750908|202 ···················· ···················· ···················· ···················· ···················· ····················
AA 6767 hCoV-19/Denmark/DCGC-17326/2020|EPI_ISL_750909|202 ···················· ···················· ···················· ···················· ···················· ····················
AA 6768 hCoV-19/Denmark/DCGC-17372/2020|EPI_ISL_750940|202 ···················· ···················· ···················· ···················· ···················· ····················
AA 6769 hCoV-19/Denmark/DCGC-17378/2020|EPI_ISL_750941|202 ···················· ···················· ···················· ···················· ···················· ····················
AA 6770 hCoV-19/Denmark/DCGC-17379/2020|EPI_ISL_750942|202 ···················· ·············A······ ···················· ···················· ···················· ····················
AA 6771 hCoV-19/Denmark/DCGC-17381/2020|EPI_ISL_750943|202 ···················· ···················· ···················· ···················· ···················· ····················
AA 6772 hCoV-19/Denmark/DCGC-17382/2020|EPI_ISL_750944|202 ···················· ···················· ···················· ···················· ···················· ····················
AA 6773 hCoV-19/Denmark/DCGC-17383/2020|EPI_ISL_750945|202 ···················· ···················· ···················· ···················· ···················· ····················
AA 6774 hCoV-19/Denmark/DCGC-17384/2020|EPI_ISL_750946|202 ···················· ···················· ···················· ···················· ···················· ····················
AA 6775 hCoV-19/Denmark/DCGC-17385/2020|EPI_ISL_750947|202 ···················· ···················· ···················· ···················· ···················· ····················
AA 6776 hCoV-19/Denmark/DCGC-17386/2020|EPI_ISL_750948|202 ···················· ···················· ···················· ···················· ···················· ····················
AA 6777 hCoV-19/Denmark/DCGC-17389/2020|EPI_ISL_750949|202 ···················· ···················· ···················· ···················· ···················· ····················
AA 6778 hCoV-19/Denmark/DCGC-17390/2020|EPI_ISL_750950|202 ···················· ···················· ···················· ···················· ···················· ····················
AA 6779 hCoV-19/Denmark/DCGC-17391/2020|EPI_ISL_750951|202 ···················· ···················· ···················· ···················· ···················· ····················
AA 6780 hCoV-19/Denmark/DCGC-17392/2020|EPI_ISL_750952|202 ···················· ·A·················· ···················· ···················· ···················· ····················
AA 6781 hCoV-19/Denmark/DCGC-17393/2020|EPI_ISL_750953|202 ···················· ···················· ···················· ···················· ···················· ····················
AA 6782 hCoV-19/Denmark/DCGC-17394/2020|EPI_ISL_750954|202 ···················· ···················· ···················· ···················· ···················· ····················
AA 6783 hCoV-19/Denmark/DCGC-17395/2020|EPI_ISL_750955|202 ···················· ···················· ···················· ···················· ···················· ····················
AA 6784 hCoV-19/Denmark/DCGC-17398/2020|EPI_ISL_750956|202 ···················· ···················· ···················· ···················· ···················· ····················
AA 6785 hCoV-19/Denmark/DCGC-17401/2020|EPI_ISL_750957|202 ···················· ···················· ···················· ···················· ···················· ····················
AA 6786 hCoV-19/Denmark/DCGC-17402/2020|EPI_ISL_750958|202 ···················· ···················· ···················· ···················· ···················· ····················
AA 6787 hCoV-19/Denmark/DCGC-17405/2020|EPI_ISL_750959|202 ···················· ···················· ···················· ···················· ···················· ····················
AA 6788 hCoV-19/Denmark/DCGC-17406/2020|EPI_ISL_750960|202 ···················· ···················· ···················· ···················· ···················· ····················
AA 6789 hCoV-19/Denmark/DCGC-17409/2020|EPI_ISL_750961|202 ···················· ···················· ···················· ···················· ···················· ····················
AA 6790 hCoV-19/Denmark/DCGC-17410/2020|EPI_ISL_750962|202 ···················· ···················· ···················· ···················· ···················· ····················
AA 6791 hCoV-19/Denmark/DCGC-17412/2020|EPI_ISL_750963|202 ···················· ·A·················· ···················· ···················· ···················· ··A·················
AA 6792 hCoV-19/Denmark/DCGC-17413/2020|EPI_ISL_750964|202 ···················· ···················· ···················· ···················· ···················· ····················
AA 6793 hCoV-19/Denmark/DCGC-17414/2020|EPI_ISL_750965|202 ···················· ···················· ···················· ···················· ···················· ····················
AA 6794 hCoV-19/Denmark/DCGC-17415/2020|EPI_ISL_750966|202 ···················· ···················· ···················· ···················· ···················· ····················
AA 6795 hCoV-19/Denmark/DCGC-17417/2020|EPI_ISL_750967|202 ···················· ·A·················· ···················· ···················· ···················· ··A·················
AA 6796 hCoV-19/Denmark/DCGC-17418/2020|EPI_ISL_750968|202 ···················· ···················· ···················· ···················· ···················· ····················
AA 6797 hCoV-19/Denmark/DCGC-17419/2020|EPI_ISL_750969|202 ···················· ···················· ···················· ···················· ···················· ····················
AA 6798 hCoV-19/Denmark/DCGC-17421/2020|EPI_ISL_750970|202 ···················· ···················· ···················· ···················· ···················· ····················
AA 6799 hCoV-19/Denmark/DCGC-17422/2020|EPI_ISL_750971|202 ···················· ···················· ···················· ···················· ······T············· ····················
AA 6800 hCoV-19/Denmark/DCGC-17425/2020|EPI_ISL_750972|202 ···················· ···················· ···················· ···················· ···················· ····················
AA 6801 hCoV-19/Denmark/DCGC-17426/2020|EPI_ISL_750973|202 ···················· ···················· ···················· ···················· ···················· ····················
AA 6802 hCoV-19/Denmark/DCGC-17427/2020|EPI_ISL_750974|202 ···················· ···················· ···················· ···················· ···················· ····················
AA 6803 hCoV-19/Denmark/DCGC-17428/2020|EPI_ISL_750975|202 ···················· ···················· ···················· ···················· ···················· ····················
AA 6804 hCoV-19/Denmark/DCGC-17429/2020|EPI_ISL_750976|202 ···················· ···················· ···················· ···················· ···················· ····················
AA 6805 hCoV-19/Denmark/DCGC-17430/2020|EPI_ISL_750977|202 ···················· ···················· ···················· ···················· ···················· ····················
AA 6806 hCoV-19/Denmark/DCGC-17431/2020|EPI_ISL_750978|202 ···················· ···················· ···················· ···················· ······T············· ····················
AA 6807 hCoV-19/Denmark/DCGC-17433/2020|EPI_ISL_750979|202 ···················· ···················· ···················· ···················· ···················· ····················
AA 6808 hCoV-19/Denmark/DCGC-17434/2020|EPI_ISL_750980|202 ···················· ···················· ···················· ···················· ···················· ····················
AA 6809 hCoV-19/Denmark/DCGC-17435/2020|EPI_ISL_750981|202 ···················· ···················· ···················· ···················· ···················· ····················
AA 6810 hCoV-19/Denmark/DCGC-17438/2020|EPI_ISL_750982|202 ···················· ···················· ···················· ···················· ···················· ····················
AA 6811 hCoV-19/USA/HI-H200579/2020|EPI_ISL_753106|2020-10 ···················· ·A·················· ···················· ···················· ···················· ····················
AA 6812 hCoV-19/USA/HI-H200580/2020|EPI_ISL_753107|2020-10 ···················· ·A·················· ···················· ···················· ···················· ····················
AA 6813 hCoV-19/USA/HI-H200582/2020|EPI_ISL_753108|2020-10 ···················· ·A·················· ···················· ···················· ···················· ····················
AA 6814 hCoV-19/USA/HI-H200583/2020|EPI_ISL_753109|2020-10 ···················· ·A·················· ···················· ···················· ···················· ····················

| | | | | | | | |
|---|---|---|---|---|---|---|---|
| AA | 6815 hCoV-19/USA/HI-H200584/2020\|EPI_ISL_753110\|2020-11 | ···················· | ·A·················· | ···················· | ···················· | ···················· | ···················· |
| AA | 6816 hCoV-19/USA/HI-H200585/2020\|EPI_ISL_753111\|2020-11 | ···················· | ·A·················· | ···················· | ···················· | ···················· | ···················· |
| AA | 6817 hCoV-19/USA/HI-H200586/2020\|EPI_ISL_753112\|2020-11 | ···················· | ·A·················· | ···················· | ···················· | ···················· | ···················· |
| AA | 6818 hCoV-19/USA/HI-H200587/2020\|EPI_ISL_753113\|2020-11 | ···················· | ·A·················· | ···················· | ···················· | ···················· | ···················· |
| AA | 6819 hCoV-19/USA/HI-H200588/2020\|EPI_ISL_753114\|2020-11 | ···················· | ·A·················· | ···················· | ···················· | ···················· | ···················· |
| AA | 6820 hCoV-19/USA/HI-H200589/2020\|EPI_ISL_753115\|2020-11 | ···················· | ·A·················· | ···················· | ···················· | ···················· | ···················· |
| AA | 6821 hCoV-19/USA/HI-H200590/2020\|EPI_ISL_753116\|2020-11 | ···················· | ···················· | ···················· | ···················· | ···················· | ···················· |
| AA | 6822 hCoV-19/USA/HI-H200591/2020\|EPI_ISL_753117\|2020-11 | ···················· | ·A·················· | ···················· | ···················· | ···················· | ···················· |
| AA | 6823 hCoV-19/USA/HI-H200592/2020\|EPI_ISL_753118\|2020-11 | ···················· | ·A·················· | ···················· | ···················· | ···················· | ···················· |
| AA | 6824 hCoV-19/USA/HI-H200593/2020\|EPI_ISL_753119\|2020-11 | ···················· | ·A·················· | ···················· | ···················· | ···················· | ···················· |
| AA | 6825 hCoV-19/USA/HI-H200594/2020\|EPI_ISL_753120\|2020-11 | ···················· | ·A·················· | ···················· | ···················· | ···················· | ···················· |
| AA | 6826 hCoV-19/USA/HI-H200595/2020\|EPI_ISL_753121\|2020-11 | ···················· | ·A·················· | ···················· | ···················· | ···················· | ···················· |
| AA | 6827 hCoV-19/USA/HI-H200596/2020\|EPI_ISL_753122\|2020-11 | ···················· | ·A·················· | ···················· | ···················· | ···················· | ···················· |
| AA | 6828 hCoV-19/USA/HI-H200597/2020\|EPI_ISL_753123\|2020-09 | ···················· | ·A·················· | ···················· | ···················· | ···················· | ···················· |
| AA | 6829 hCoV-19/USA/HI-H200598/2020\|EPI_ISL_753124\|2020-10 | ···················· | ·A·················· | ···················· | ···················· | ···················· | ···················· |
| AA | 6830 hCoV-19/USA/HI-H200599/2020\|EPI_ISL_753125\|2020-10 | ···················· | ·A·················· | ···················· | ···················· | ···················· | ···················· |
| AA | 6831 hCoV-19/USA/HI-H200600/2020\|EPI_ISL_753126\|2020-10 | ···················· | ·A·················· | ···················· | ···················· | ···················· | ···················· |
| AA | 6832 hCoV-19/USA/HI-H200601/2020\|EPI_ISL_753127\|2020-10 | ···················· | ·A·················· | ···················· | ···················· | ···················· | ···················· |
| AA | 6833 hCoV-19/USA/HI-H200602/2020\|EPI_ISL_753128\|2020-10 | ···················· | ·A·················· | ···················· | ···················· | ···················· | ···················· |
| AA | 6834 hCoV-19/USA/HI-H200603/2020\|EPI_ISL_753129\|2020-11 | ···················· | ·A·················· | ···················· | ···················· | ···················· | ···················· |
| AA | 6835 hCoV-19/USA/HI-H200604/2020\|EPI_ISL_753130\|2020-11 | ···················· | ·A·················· | ···················· | ···················· | ···················· | ···················· |
| AA | 6836 hCoV-19/USA/HI-H200605/2020\|EPI_ISL_753131\|2020-11 | ···················· | ·A·················· | ···················· | ···················· | ···················· | ···················· |
| AA | 6837 hCoV-19/USA/HI-H200606/2020\|EPI_ISL_753132\|2020-11 | ···················· | ·A·················· | ···················· | ···················· | ···················· | ···················· |
| AA | 6838 hCoV-19/USA/HI-H200607/2020\|EPI_ISL_753133\|2020-11 | ···················· | ·A·················· | ···················· | ···················· | ···················· | ···················· |
| AA | 6839 hCoV-19/USA/HI-H200608/2020\|EPI_ISL_753134\|2020-11 | ···················· | ·A·················· | ···················· | ···················· | ···················· | ···················· |
| AA | 6840 hCoV-19/USA/HI-H200609/2020\|EPI_ISL_753135\|2020-11 | ···················· | ·A·················· | ···················· | ···················· | ···················· | ···················· |
| AA | 6841 hCoV-19/USA/HI-H200610/2020\|EPI_ISL_753136\|2020-11 | ···················· | ·A·················· | ···················· | ···················· | ···················· | ···················· |
| AA | 6842 hCoV-19/USA/HI-H200611/2020\|EPI_ISL_753137\|2020-11 | ···················· | ·A·················· | ···················· | ···················· | ···················· | ···················· |
| AA | 6843 hCoV-19/USA/HI-H200612/2020\|EPI_ISL_753138\|2020-11 | ···················· | ·A·················· | ···················· | ···················· | ···················· | ···················· |
| AA | 6844 hCoV-19/USA/NY-Wadsworth-282766-01/2020\|EPI_ISL_75 | ···················· | ···················· | ···················· | ···················· | ···················· | ···················· |
| AA | 6845 hCoV-19/USA/NY-Wadsworth-282774-01/2020\|EPI_ISL_75 | ···················· | ···················· | ···················· | ···················· | ···················· | ···················· |
| AA | 6846 hCoV-19/USA/NY-Wadsworth-282803-01/2020\|EPI_ISL_75 | ···················· | ···················· | ···················· | ···················· | ···················· | ···················· |
| AA | 6847 hCoV-19/USA/NY-Wadsworth-283201-01/2020\|EPI_ISL_75 | ···················· | ···················· | ···················· | ···················· | ···················· | ···················· |
| AA | 6848 hCoV-19/USA/NY-Wadsworth-283208-01/2020\|EPI_ISL_75 | ···················· | ···················· | ···················· | ···················· | ···················· | ···················· |
| AA | 6849 hCoV-19/USA/NY-Wadsworth-283255-01/2020\|EPI_ISL_75 | ···················· | ···················· | ···················· | ···················· | ···················· | ···················· |
| AA | 6850 hCoV-19/USA/NY-Wadsworth-283435-01/2020\|EPI_ISL_75 | ···················· | ···················· | ···················· | ···················· | ···················· | ···················· |
| AA | 6851 hCoV-19/USA/NY-Wadsworth-283448-01/2020\|EPI_ISL_75 | ···················· | ·A·················· | ···················· | ···················· | ···················· | ···················· |
| AA | 6852 hCoV-19/USA/NY-Wadsworth-283449-01/2020\|EPI_ISL_75 | ···················· | ·A·················· | ···················· | ···················· | ···················· | ···················· |
| AA | 6853 hCoV-19/USA/NY-Wadsworth-283453-01/2020\|EPI_ISL_75 | ···················· | ···················· | ···················· | ···················· | ···················· | ···················· |
| AA | 6854 hCoV-19/USA/NY-Wadsworth-283483-01/2020\|EPI_ISL_75 | ···················· | ···················· | ···················· | ···················· | ···················· | ···················· |
| AA | 6855 hCoV-19/USA/NY-Wadsworth-283661-01/2020\|EPI_ISL_75 | ···················· | ···················· | ···················· | ···················· | ···················· | ···················· |
| AA | 6856 hCoV-19/USA/NY-Wadsworth-283604-01/2020\|EPI_ISL_75 | ···················· | ·A·················· | ···················· | ···················· | ···················· | ···················· |
| AA | 6857 hCoV-19/USA/NY-Wadsworth-283777-01/2020\|EPI_ISL_75 | ···················· | ·A·················· | ···················· | ···················· | ···················· | ···················· |
| AA | 6858 hCoV-19/USA/NY-Wadsworth-283767-01/2020\|EPI_ISL_75 | ···················· | ···················· | ···················· | ···················· | ···················· | ···················· |
| AA | 6859 hCoV-19/USA/NY-Wadsworth-284180-01/2020\|EPI_ISL_75 | ···················· | ···················· | ···················· | ···················· | ···················· | ···················· |
| AA | 6860 hCoV-19/USA/NY-Wadsworth-284741-01/2020\|EPI_ISL_75 | ···················· | ·A·················· | ···················· | ···················· | ···················· | ···················· |
| AA | 6861 hCoV-19/USA/NY-Wadsworth-283760-01/2020\|EPI_ISL_75 | ···················· | ·A·················· | ···················· | ···················· | ···················· | ···················· |
| AA | 6862 hCoV-19/USA/NY-Wadsworth-284730-01/2020\|EPI_ISL_75 | ···················· | ···················· | ···················· | ···················· | ···················· | ···················· |
| AA | 6863 hCoV-19/USA/NY-Wadsworth-284731-01/2020\|EPI_ISL_75 | ···················· | ···················· | ···················· | ···················· | ···················· | ···················· |
| AA | 6864 hCoV-19/USA/NY-Wadsworth-284737-01/2020\|EPI_ISL_75 | ···················· | ···················· | ···················· | ···················· | ···················· | ···················· |
| AA | 6865 hCoV-19/USA/NY-Wadsworth-284724-01/2020\|EPI_ISL_75 | ···················· | ···················· | ···················· | ···················· | ···················· | ···················· |
| AA | 6866 hCoV-19/USA/NY-Wadsworth-284794-01/2020\|EPI_ISL_75 | ···················· | ···················· | ···················· | ···················· | ···················· | ···················· |
| AA | 6867 hCoV-19/USA/NY-Wadsworth-284575-01/2020\|EPI_ISL_75 | ···················· | ···················· | ···················· | ···················· | ···················· | ···················· |
| AA | 6868 hCoV-19/USA/NY-Wadsworth-285144-01/2020\|EPI_ISL_75 | ···················· | ···················· | ···················· | ···················· | ···················· | ···················· |
| AA | 6869 hCoV-19/USA/NY-Wadsworth-285193-01/2020\|EPI_ISL_75 | ···················· | ···················· | ···················· | ···················· | ···················· | ···················· |
| AA | 6870 hCoV-19/USA/NY-Wadsworth-285191-01/2020\|EPI_ISL_75 | ···················· | ···················· | ···················· | ···················· | ···················· | ···················· |
| AA | 6871 hCoV-19/USA/NY-Wadsworth-285166-01/2020\|EPI_ISL_75 | ···················· | ···················· | ···················· | ···················· | ···················· | ···················· |
| AA | 6872 hCoV-19/USA/NY-Wadsworth-285156-01/2020\|EPI_ISL_75 | ···················· | ···················· | ···················· | ···················· | ···················· | ···················· |
| AA | 6873 hCoV-19/USA/NY-Wadsworth-285155-01/2020\|EPI_ISL_75 | ···················· | ···················· | ···················· | ···················· | ···················· | ···················· |
| AA | 6874 hCoV-19/USA/NY-Wadsworth-285154-01/2020\|EPI_ISL_75 | ···················· | ···················· | ···················· | ···················· | ···················· | ···················· |
| AA | 6875 hCoV-19/USA/NY-Wadsworth-285009-01/2020\|EPI_ISL_75 | ···················· | ···················· | ···················· | ···················· | ···················· | ···················· |
| AA | 6876 hCoV-19/USA/NY-Wadsworth-285002-01/2020\|EPI_ISL_75 | ···················· | ···················· | ···················· | ···················· | ···················· | ···················· |
| AA | 6877 hCoV-19/Canada/ON-S1767/2020\|EPI_ISL_755768\|2020-0 | ···················· | ···················· | ···················· | ···················· | ···················· | ···················· |
| AA | 6878 hCoV-19/Canada/ON-S1771/2020\|EPI_ISL_755772\|2020-0 | ···················· | ···················· | ···················· | ···················· | ···················· | ···················· |
| AA | 6879 hCoV-19/Canada/ON-S1774/2020\|EPI_ISL_755774\|2020-0 | ···················· | ···················· | ···················· | ···················· | ···················· | ···················· |
| AA | 6880 hCoV-19/Canada/ON-S1775/2020\|EPI_ISL_755775\|2020-0 | ···················· | ···················· | ···················· | ···················· | ···················· | ···················· |
| AA | 6881 hCoV-19/Canada/ON-S1778/2020\|EPI_ISL_755776\|2020-0 | ···················· | ···················· | ···················· | ···················· | ···················· | ···················· |
| AA | 6882 hCoV-19/Canada/ON-S1779/2020\|EPI_ISL_755777\|2020-0 | ···················· | ···················· | ···················· | ···················· | ···················· | ···················· |
| AA | 6883 hCoV-19/Canada/ON-S1780/2020\|EPI_ISL_755778\|2020-0 | ···················· | ·A·················· | ···················· | ···················· | ···················· | ··A················· |
| AA | 6884 hCoV-19/Canada/ON-S1781/2020\|EPI_ISL_755779\|2020-0 | ···················· | ···················· | ···················· | ···················· | ···················· | ···················· |
| AA | 6885 hCoV-19/Canada/ON-S1783/2020\|EPI_ISL_755781\|2020-0 | ···················· | ···················· | ···················· | ···················· | ···················· | ···················· |
| AA | 6886 hCoV-19/Canada/ON-S1786/2020\|EPI_ISL_755783\|2020-1 | ···················· | ···················· | ···················· | ···················· | ···················· | ···················· |
| AA | 6887 hCoV-19/Canada/ON-S1789/2020\|EPI_ISL_755785\|2020-1 | ···················· | ·A·················· | ···················· | ···················· | ···················· | ··A················· |
| AA | 6888 hCoV-19/Canada/ON-S1790/2020\|EPI_ISL_755786\|2020-1 | ···················· | ···················· | ···················· | ···················· | ···················· | ···················· |
| AA | 6889 hCoV-19/Canada/ON-S1793/2020\|EPI_ISL_755788\|2020-1 | ···················· | ···················· | ···················· | ···················· | ···················· | ···················· |
| AA | 6890 hCoV-19/Canada/ON-S1795/2020\|EPI_ISL_755790\|2020-1 | ···················· | ···················· | ···················· | ···················· | ···················· | ···················· |
| AA | 6891 hCoV-19/Canada/ON-S1797/2020\|EPI_ISL_755792\|2020-1 | ···················· | ·A·················· | ···················· | ···················· | ···················· | ··A················· |
| AA | 6892 hCoV-19/Canada/ON-S193/2020\|EPI_ISL_755796\|2020-03 | ···················· | ···················· | ···················· | ···················· | ···················· | ···················· |
| AA | 6893 hCoV-19/Denmark/DCGC-20686/2020\|EPI_ISL_759300\|202 | ···················· | ···················· | ···················· | ···················· | ···················· | ···················· |
| AA | 6894 hCoV-19/Denmark/DCGC-20687/2020\|EPI_ISL_759301\|202 | ···················· | ···················· | ···················· | ···················· | ···················· | ···················· |

| | | | | | | | | |
|---|---|---|---|---|---|---|---|---|
| AA | 6895 | hCoV-19/Denmark/DCGC-20688/2020\|EPI_ISL_759302\|202 | ···················· | ····A··············· | ···················· | ···················· | ···················· | ···················· |
| AA | 6896 | hCoV-19/Denmark/DCGC-20689/2020\|EPI_ISL_759303\|202 | ···················· | ···················· | ···················· | ···················· | ···················· | ···················· |
| AA | 6897 | hCoV-19/Denmark/DCGC-20691/2020\|EPI_ISL_759304\|202 | ···················· | ···················· | ···················· | ···················· | ···················· | ···················· |
| AA | 6898 | hCoV-19/Denmark/DCGC-20693/2020\|EPI_ISL_759305\|202 | ···················· | ···················· | ···················· | ···················· | ···················· | ···················· |
| AA | 6899 | hCoV-19/Denmark/DCGC-20695/2020\|EPI_ISL_759306\|202 | ···················· | ···················· | ···················· | ···················· | ···················· | ···················· |
| AA | 6900 | hCoV-19/Denmark/DCGC-20696/2020\|EPI_ISL_759307\|202 | ···················· | ···················· | ···················· | ···················· | ···················· | ···················· |
| AA | 6901 | hCoV-19/Denmark/DCGC-20697/2020\|EPI_ISL_759308\|202 | ···················· | ···················· | ···················· | ···················· | ···················· | ···················· |
| AA | 6902 | hCoV-19/Denmark/DCGC-20698/2020\|EPI_ISL_759309\|202 | ···················· | ···················· | ···················· | ···················· | ···················· | ···················· |
| AA | 6903 | hCoV-19/Denmark/DCGC-20699/2020\|EPI_ISL_759310\|202 | ···················· | ···················· | ···················· | ···················· | ···················· | ···················· |
| AA | 6904 | hCoV-19/Denmark/DCGC-20700/2020\|EPI_ISL_759311\|202 | ···················· | ···················· | ···················· | ···················· | ···················· | ···················· |
| AA | 6905 | hCoV-19/Denmark/DCGC-20701/2020\|EPI_ISL_759312\|202 | ···················· | ···················· | ···················· | ···················· | ···················· | ···················· |
| AA | 6906 | hCoV-19/Denmark/DCGC-20702/2020\|EPI_ISL_759313\|202 | ···················· | ···················· | ···················· | ···················· | ···················· | ···················· |
| AA | 6907 | hCoV-19/Denmark/DCGC-20703/2020\|EPI_ISL_759314\|202 | ···················· | ·A·················· | ···················· | ···················· | ···················· | ···················· |
| AA | 6908 | hCoV-19/Denmark/DCGC-20704/2020\|EPI_ISL_759315\|202 | ···················· | ···················· | ···················· | ···················· | ···················· | ···················· |
| AA | 6909 | hCoV-19/Denmark/DCGC-20705/2020\|EPI_ISL_759316\|202 | ···················· | ···················· | ···················· | ···················· | ···················· | ···················· |
| AA | 6910 | hCoV-19/Denmark/DCGC-20708/2020\|EPI_ISL_759317\|202 | ···················· | ···················· | ···················· | ···················· | ···················· | ···················· |
| AA | 6911 | hCoV-19/Denmark/DCGC-20709/2020\|EPI_ISL_759318\|202 | ···················· | ···················· | ···················· | ···················· | ···················· | ···················· |
| AA | 6912 | hCoV-19/Denmark/DCGC-20710/2020\|EPI_ISL_759319\|202 | ···················· | ···················· | ···················· | ···················· | ···················· | ···················· |
| AA | 6913 | hCoV-19/South Korea/KDCA0059/2020\|EPI_ISL_747300\|2 | ···················· | ···················· | ···················· | ···················· | ···················· | ···················· |
| AA | 6914 | hCoV-19/South Korea/KDCA0060/2020\|EPI_ISL_747301\|2 | ···················· | ···················· | ···················· | ···················· | ···················· | ···················· |
| AA | 6915 | hCoV-19/South Korea/KDCA0061/2020\|EPI_ISL_747302\|2 | ···················· | ···················· | ···················· | ···················· | ···················· | ···················· |
| AA | 6916 | hCoV-19/South Korea/KDCA0062/2020\|EPI_ISL_747303\|2 | ···················· | ···················· | ···················· | ···················· | ···················· | ···················· |
| AA | 6917 | hCoV-19/South Korea/KDCA0065/2020\|EPI_ISL_747306\|2 | ···················· | ···················· | ···················· | ···················· | ···················· | ···················· |
| AA | 6918 | hCoV-19/South Korea/KDCA0067/2020\|EPI_ISL_747308\|2 | ···················· | ···················· | ···················· | ···················· | ···················· | ···················· |
| AA | 6919 | hCoV-19/South Korea/KDCA0068/2020\|EPI_ISL_747309\|2 | ···················· | ···················· | ···················· | ···················· | ···················· | ···················· |
| AA | 6920 | hCoV-19/South Korea/KDCA0069/2020\|EPI_ISL_747310\|2 | ···················· | ···················· | ···················· | ···················· | ···················· | ···················· |
| AA | 6921 | hCoV-19/South Korea/KDCA0071/2020\|EPI_ISL_747312\|2 | ···················· | ···················· | ···················· | ···················· | ···················· | ···················· |
| AA | 6922 | hCoV-19/South Korea/KDCA0072/2020\|EPI_ISL_747313\|2 | ···················· | ···················· | ···················· | ···················· | ···················· | ···················· |
| AA | 6923 | hCoV-19/South Korea/KDCA0073/2020\|EPI_ISL_747314\|2 | ···················· | ···················· | ···················· | ···················· | ···················· | ···················· |
| AA | 6924 | hCoV-19/South Korea/KDCA0074/2020\|EPI_ISL_747315\|2 | ···················· | ···················· | ···················· | ···················· | ···················· | ···················· |
| AA | 6925 | hCoV-19/South Korea/KDCA0075/2020\|EPI_ISL_747316\|2 | ···················· | ···················· | ···················· | ···················· | ···················· | ···················· |
| AA | 6926 | hCoV-19/South Korea/KDCA0077/2020\|EPI_ISL_747317\|2 | ···················· | ···················· | ···················· | ···················· | ···················· | ···················· |
| AA | 6927 | hCoV-19/South Korea/KDCA0079/2020\|EPI_ISL_747319\|2 | ···················· | ···················· | ···················· | ···················· | ···················· | ···················· |
| AA | 6928 | hCoV-19/South Korea/KDCA0080/2020\|EPI_ISL_747320\|2 | ···················· | ···················· | ···················· | ···················· | ···················· | ···················· |
| AA | 6929 | hCoV-19/South Korea/KDCA0081/2020\|EPI_ISL_747321\|2 | ···················· | ···················· | ···················· | ···················· | ···················· | ···················· |
| AA | 6930 | hCoV-19/South Korea/KDCA0082/2020\|EPI_ISL_747322\|2 | ···················· | ···················· | ···················· | ···················· | ···················· | ···················· |
| AA | 6931 | hCoV-19/South Korea/KDCA0085/2020\|EPI_ISL_747324\|2 | ···················· | ···················· | ···················· | ···················· | ···················· | ···················· |
| AA | 6932 | hCoV-19/South Korea/KDCA0086/2020\|EPI_ISL_747325\|2 | ···················· | ···················· | ···················· | ···················· | ···················· | ···················· |
| AA | 6933 | hCoV-19/South Korea/KDCA0087/2020\|EPI_ISL_747326\|2 | ···················· | ···················· | ···················· | ···················· | ···················· | ···················· |
| AA | 6934 | hCoV-19/South Korea/KDCA0088/2020\|EPI_ISL_747327\|2 | ···················· | ···················· | ···················· | ···················· | ···················· | ···················· |
| AA | 6935 | hCoV-19/South Korea/KDCA0089/2020\|EPI_ISL_747328\|2 | ···················· | ···················· | ···················· | ···················· | ···················· | ···················· |
| AA | 6936 | hCoV-19/South Korea/KDCA0090/2020\|EPI_ISL_747329\|2 | ···················· | ···················· | ···················· | ···················· | ···················· | ···················· |
| AA | 6937 | hCoV-19/South Korea/KDCA0091/2020\|EPI_ISL_747330\|2 | ···················· | ···················· | ···················· | ···················· | ···················· | ···················· |
| AA | 6938 | hCoV-19/South Korea/KDCA0109/2020\|EPI_ISL_747345\|2 | ···················· | ·A·················· | ···················· | ···················· | ···················· | ···A················ |
| AA | 6939 | hCoV-19/South Korea/KDCA0111/2020\|EPI_ISL_747346\|2 | ···················· | ···················· | ···················· | ···················· | ···················· | ···················· |
| AA | 6940 | hCoV-19/South Korea/KDCA0112/2020\|EPI_ISL_747347\|2 | ···················· | ···················· | ···················· | ···················· | ···················· | ···················· |
| AA | 6941 | hCoV-19/South Korea/KDCA0115/2020\|EPI_ISL_747350\|2 | ···················· | ···················· | ···················· | ···················· | ···················· | ···················· |
| AA | 6942 | hCoV-19/South Korea/KDCA0116/2020\|EPI_ISL_747351\|2 | ···················· | ···················· | ···················· | ···················· | ···················· | ···················· |
| AA | 6943 | hCoV-19/South Korea/KDCA0117/2020\|EPI_ISL_747352\|2 | ···················· | ···················· | ···················· | ···················· | ···················· | ···················· |
| AA | 6944 | hCoV-19/South Korea/KDCA0118/2020\|EPI_ISL_747353\|2 | ···················· | ···················· | ···················· | ···················· | ···················· | ···················· |
| AA | 6945 | hCoV-19/South Korea/KDCA0120/2020\|EPI_ISL_747355\|2 | ···················· | ···················· | ···················· | ···················· | ···················· | ···················· |
| AA | 6946 | hCoV-19/South Korea/KDCA0121/2020\|EPI_ISL_747356\|2 | ···················· | ···················· | ···················· | ···················· | ···················· | ···················· |
| AA | 6947 | hCoV-19/South Korea/KDCA0122/2020\|EPI_ISL_747357\|2 | ···················· | ···················· | ···················· | ···················· | ···················· | ···················· |
| AA | 6948 | hCoV-19/South Korea/KDCA0123/2020\|EPI_ISL_747358\|2 | ···················· | ···················· | ···················· | ···················· | ···················· | ···················· |
| AA | 6949 | hCoV-19/South Korea/KDCA0124/2020\|EPI_ISL_747359\|2 | ···················· | ···················· | ···················· | ···················· | ···················· | ···················· |
| AA | 6950 | hCoV-19/South Korea/KDCA0125/2020\|EPI_ISL_747360\|2 | ···················· | ···················· | ···················· | ···················· | ···················· | ···················· |
| AA | 6951 | hCoV-19/South Korea/KDCA0126/2020\|EPI_ISL_747361\|2 | ···················· | ···················· | ···················· | ···················· | ···················· | ···················· |
| AA | 6952 | hCoV-19/South Korea/KDCA0128/2020\|EPI_ISL_747363\|2 | ···················· | ···················· | ···················· | ···················· | ···················· | ···················· |
| AA | 6953 | hCoV-19/South Korea/KDCA0129/2020\|EPI_ISL_747364\|2 | ···················· | ···················· | ···················· | ···················· | ···················· | ···················· |
| AA | 6954 | hCoV-19/South Korea/KDCA0130/2020\|EPI_ISL_747365\|2 | ···················· | ···················· | ···················· | ···················· | ···················· | ···················· |
| AA | 6955 | hCoV-19/South Korea/KDCA0131/2020\|EPI_ISL_747366\|2 | ···················· | ···················· | ···················· | ···················· | ···················· | ···A················ |
| AA | 6956 | hCoV-19/South Korea/KDCA0132/2020\|EPI_ISL_747367\|2 | ···················· | ···················· | ···················· | ···················· | ···················· | ···················· |
| AA | 6957 | hCoV-19/South Korea/KDCA0133/2020\|EPI_ISL_747368\|2 | ···················· | ···················· | ···················· | ···················· | ···················· | ···················· |
| AA | 6958 | hCoV-19/South Korea/KDCA0134/2020\|EPI_ISL_747369\|2 | ···················· | ···················· | ···················· | ···················· | ···················· | ···················· |
| AA | 6959 | hCoV-19/South Korea/KDCA0135/2020\|EPI_ISL_747370\|2 | ···················· | ···················· | ···················· | ···················· | ···················· | ···················· |
| AA | 6960 | hCoV-19/South Korea/KDCA0136/2020\|EPI_ISL_747371\|2 | ···················· | ···················· | ···················· | ···················· | ···················· | ···················· |
| AA | 6961 | hCoV-19/South Korea/KDCA0137/2020\|EPI_ISL_747372\|2 | ···················· | ···················· | ···················· | ···················· | ···················· | ···················· |
| AA | 6962 | hCoV-19/South Korea/KDCA0138/2020\|EPI_ISL_747373\|2 | ···················· | ···················· | ···················· | ···················· | ···················· | ···················· |
| AA | 6963 | hCoV-19/South Korea/KDCA0139/2020\|EPI_ISL_747374\|2 | ···················· | ···················· | ···················· | ···················· | ···················· | ···················· |
| AA | 6964 | hCoV-19/South Korea/KDCA0140/2020\|EPI_ISL_747375\|2 | ···················· | ···················· | ···················· | ···················· | ···················· | ···················· |
| AA | 6965 | hCoV-19/South Korea/KDCA0142/2020\|EPI_ISL_747377\|2 | ···················· | ···················· | ···················· | ···················· | ···················· | ···················· |
| AA | 6966 | hCoV-19/South Korea/KDCA0143/2020\|EPI_ISL_747378\|2 | ···················· | ···················· | ···················· | ···················· | ···················· | ···················· |
| AA | 6967 | hCoV-19/South Korea/KDCA0144/2020\|EPI_ISL_747379\|2 | ···················· | ···················· | ···················· | ···················· | ···················· | ···················· |
| AA | 6968 | hCoV-19/Denmark/DCGC-18903/2020\|EPI_ISL_748629\|202 | ···················· | ···················· | ···················· | ···················· | ···················· | ···················· |
| AA | 6969 | hCoV-19/Denmark/DCGC-18904/2020\|EPI_ISL_748630\|202 | ···················· | ···················· | ···················· | ···················· | ···················· | ···················· |
| AA | 6970 | hCoV-19/Denmark/DCGC-18906/2020\|EPI_ISL_748631\|202 | ···················· | ···················· | ···················· | ···················· | ···················· | ···················· |
| AA | 6971 | hCoV-19/Denmark/DCGC-18907/2020\|EPI_ISL_748632\|202 | ···················· | ···················· | ···················· | ···················· | ···················· | ···················· |
| AA | 6972 | hCoV-19/Denmark/DCGC-18908/2020\|EPI_ISL_748633\|202 | ···················· | ···················· | ···················· | ···················· | ···················· | ···················· |
| AA | 6973 | hCoV-19/Denmark/DCGC-18909/2020\|EPI_ISL_748634\|202 | ···················· | ···················· | ···················· | ···················· | ···················· | ···················· |
| AA | 6974 | hCoV-19/Denmark/DCGC-18910/2020\|EPI_ISL_748635\|202 | ···················· | ···················· | ···················· | ···················· | ···················· | ···················· |

| | | | | | | | | |
|---|---|---|---|---|---|---|---|---|
| AA | 6975 | hCoV-19/Denmark/DCGC-18912/2020\|EPI_ISL_748636\|202 | ···················· | ···················· | ···················· | ···················· | ···················· | ···················· |
| AA | 6976 | hCoV-19/Denmark/DCGC-18913/2020\|EPI_ISL_748637\|202 | ···················· | ···················· | ···················· | ···················· | ···················· | ···················· |
| AA | 6977 | hCoV-19/Denmark/DCGC-18914/2020\|EPI_ISL_748638\|202 | ···················· | ···················· | ···················· | ···················· | ···················· | ···················· |
| AA | 6978 | hCoV-19/Denmark/DCGC-18915/2020\|EPI_ISL_748639\|202 | ···················· | ···················· | ···················· | ···················· | ···················· | ···················· |
| AA | 6979 | hCoV-19/Denmark/DCGC-18920/2020\|EPI_ISL_748640\|202 | ···················· | ···················· | ···················· | ···················· | ···················· | ···················· |
| AA | 6980 | hCoV-19/Denmark/DCGC-18921/2020\|EPI_ISL_748641\|202 | ···················· | ·A·················· | ···················· | ···················· | ···················· | ···················· |
| AA | 6981 | hCoV-19/Denmark/DCGC-18924/2020\|EPI_ISL_748642\|202 | ···················· | ···················· | ···················· | ···················· | ···················· | ···················· |
| AA | 6982 | hCoV-19/Denmark/DCGC-18926/2020\|EPI_ISL_748643\|202 | ···················· | ···················· | ···················· | ···················· | ···················· | ···················· |
| AA | 6983 | hCoV-19/Denmark/DCGC-18927/2020\|EPI_ISL_748644\|202 | ···················· | ···················· | ···················· | ···················· | ···················· | ···················· |
| AA | 6984 | hCoV-19/Denmark/DCGC-18928/2020\|EPI_ISL_748645\|202 | ···················· | ···················· | ···················· | ···················· | ···················· | ···················· |
| AA | 6985 | hCoV-19/Denmark/DCGC-18929/2020\|EPI_ISL_748646\|202 | ···················· | ···················· | ···················· | ···················· | ···················· | ···················· |
| AA | 6986 | hCoV-19/Denmark/DCGC-18930/2020\|EPI_ISL_748647\|202 | ···················· | ···················· | ···················· | ···················· | ···················· | ···················· |
| AA | 6987 | hCoV-19/Denmark/DCGC-18932/2020\|EPI_ISL_748648\|202 | ···················· | ···················· | ···················· | ···················· | ···················· | ···················· |
| AA | 6988 | hCoV-19/Denmark/DCGC-18935/2020\|EPI_ISL_748649\|202 | ···················· | ···················· | ···················· | ···················· | ···················· | ···················· |
| AA | 6989 | hCoV-19/Denmark/DCGC-18937/2020\|EPI_ISL_748650\|202 | ···················· | ···················· | ···················· | ···················· | ···················· | ···················· |
| AA | 6990 | hCoV-19/Denmark/DCGC-18938/2020\|EPI_ISL_748651\|202 | ···················· | ···················· | ···················· | ···················· | ···················· | ···················· |
| AA | 6991 | hCoV-19/Denmark/DCGC-18940/2020\|EPI_ISL_748652\|202 | ···················· | ···················· | ···················· | ···················· | ···················· | ···················· |
| AA | 6992 | hCoV-19/Denmark/DCGC-18941/2020\|EPI_ISL_748653\|202 | ···················· | ···················· | ···················· | ···················· | ···················· | ···················· |
| AA | 6993 | hCoV-19/Denmark/DCGC-18943/2020\|EPI_ISL_748654\|202 | ···················· | ···················· | ···················· | ···················· | ···················· | ···················· |
| AA | 6994 | hCoV-19/Denmark/DCGC-18944/2020\|EPI_ISL_748655\|202 | ···················· | ···················· | ···················· | ···················· | ···················· | ···················· |
| AA | 6995 | hCoV-19/Denmark/DCGC-18945/2020\|EPI_ISL_748656\|202 | ···················· | ···················· | ···················· | ···················· | ···················· | ···················· |
| AA | 6996 | hCoV-19/Denmark/DCGC-18946/2020\|EPI_ISL_748657\|202 | ···················· | ···················· | ···················· | ···················· | ···················· | ···················· |
| AA | 6997 | hCoV-19/Denmark/DCGC-18948/2020\|EPI_ISL_748658\|202 | ···················· | ···················· | ···················· | ···················· | ···················· | ···················· |
| AA | 6998 | hCoV-19/Denmark/DCGC-18949/2020\|EPI_ISL_748659\|202 | ···················· | ···················· | ···················· | ···················· | ···················· | ···················· |
| AA | 6999 | hCoV-19/Denmark/DCGC-18951/2020\|EPI_ISL_748660\|202 | ···················· | ···················· | ···················· | ···················· | ···················· | ···················· |
| AA | 7000 | hCoV-19/Denmark/DCGC-18974/2020\|EPI_ISL_748676\|202 | ···················· | ···················· | ···················· | ···················· | ···················· | ···················· |
| AA | 7001 | hCoV-19/Denmark/DCGC-18975/2020\|EPI_ISL_748677\|202 | ···················· | ···················· | ···················· | ···················· | ···················· | ···················· |
| AA | 7002 | hCoV-19/Denmark/DCGC-18976/2020\|EPI_ISL_748678\|202 | ···················· | ···················· | ···················· | ···················· | ···················· | ···················· |
| AA | 7003 | hCoV-19/Denmark/DCGC-18979/2020\|EPI_ISL_748679\|202 | ···················· | ···················· | ···················· | ···················· | ···················· | ···················· |
| AA | 7004 | hCoV-19/Denmark/DCGC-18980/2020\|EPI_ISL_748680\|202 | ···················· | ·A·················· | ···················· | ···················· | ···················· | ··A················· |
| AA | 7005 | hCoV-19/Denmark/DCGC-18982/2020\|EPI_ISL_748681\|202 | ···················· | ···················· | ···················· | ···················· | ···················· | ···················· |
| AA | 7006 | hCoV-19/Denmark/DCGC-18983/2020\|EPI_ISL_748682\|202 | ···················· | ···················· | ···················· | ···················· | ···················· | ···················· |
| AA | 7007 | hCoV-19/Denmark/DCGC-18984/2020\|EPI_ISL_748683\|202 | ···················· | ···················· | ···················· | ···················· | ···················· | ···················· |
| AA | 7008 | hCoV-19/Denmark/DCGC-18989/2020\|EPI_ISL_748684\|202 | ···················· | ···················· | ···················· | ···················· | ···················· | ···················· |
| AA | 7009 | hCoV-19/Denmark/DCGC-18990/2020\|EPI_ISL_748685\|202 | ···················· | ···················· | ···················· | ···················· | ···················· | ···················· |
| AA | 7010 | hCoV-19/Denmark/DCGC-18991/2020\|EPI_ISL_748686\|202 | ···················· | ···················· | ···················· | ···················· | ···················· | ···················· |
| AA | 7011 | hCoV-19/Denmark/DCGC-18992/2020\|EPI_ISL_748687\|202 | ···················· | ···················· | ···················· | ···················· | ···················· | ···················· |
| AA | 7012 | hCoV-19/Denmark/DCGC-18997/2020\|EPI_ISL_748688\|202 | ···················· | ·A·················· | ···················· | ···················· | ···················· | ···················· |
| AA | 7013 | hCoV-19/Denmark/DCGC-18998/2020\|EPI_ISL_748689\|202 | ···················· | ···················· | ···················· | ···················· | ···················· | ···················· |
| AA | 7014 | hCoV-19/Denmark/DCGC-19000/2020\|EPI_ISL_748690\|202 | ···················· | ···················· | ···················· | ···················· | ···················· | ···················· |
| AA | 7015 | hCoV-19/Denmark/DCGC-19001/2020\|EPI_ISL_748691\|202 | ···················· | ···················· | ···················· | ···················· | ···················· | ···················· |
| AA | 7016 | hCoV-19/Denmark/DCGC-19003/2020\|EPI_ISL_748692\|202 | ···················· | ···················· | ···················· | ···················· | ···················· | ···················· |
| AA | 7017 | hCoV-19/Denmark/DCGC-19005/2020\|EPI_ISL_748693\|202 | ···················· | ···················· | ···················· | ···················· | ···················· | ···················· |
| AA | 7018 | hCoV-19/Denmark/DCGC-19008/2020\|EPI_ISL_748694\|202 | ···················· | ···················· | ···················· | ···················· | ···················· | ···················· |
| AA | 7019 | hCoV-19/Denmark/DCGC-19010/2020\|EPI_ISL_748695\|202 | ···················· | ···················· | ···················· | ···················· | ···················· | ···················· |
| AA | 7020 | hCoV-19/Denmark/DCGC-19012/2020\|EPI_ISL_748696\|202 | ···················· | ···················· | ···················· | ···················· | ···················· | ···················· |
| AA | 7021 | hCoV-19/Denmark/DCGC-19014/2020\|EPI_ISL_748697\|202 | ···················· | ···················· | ···················· | ···················· | ···················· | ···················· |
| AA | 7022 | hCoV-19/Denmark/DCGC-19015/2020\|EPI_ISL_748698\|202 | ···················· | ···················· | ···················· | ···················· | ···················· | ···················· |
| AA | 7023 | hCoV-19/Denmark/DCGC-19016/2020\|EPI_ISL_748699\|202 | ···················· | ···················· | ···················· | ···················· | ···················· | ···················· |
| AA | 7024 | hCoV-19/Denmark/DCGC-18428/2020\|EPI_ISL_749960\|202 | ···················· | ···················· | ···················· | ···················· | ···················· | ···················· |
| AA | 7025 | hCoV-19/Denmark/DCGC-18429/2020\|EPI_ISL_749961\|202 | ···················· | ···················· | ···················· | ···················· | ···················· | ···················· |
| AA | 7026 | hCoV-19/Denmark/DCGC-18430/2020\|EPI_ISL_749962\|202 | ···················· | ···················· | ···················· | ···················· | ···················· | ···················· |
| AA | 7027 | hCoV-19/Denmark/DCGC-18431/2020\|EPI_ISL_749963\|202 | ···················· | ···················· | ···················· | ···················· | ···················· | ···················· |
| AA | 7028 | hCoV-19/Denmark/DCGC-18432/2020\|EPI_ISL_749964\|202 | ···················· | ···················· | ···················· | ···················· | ···················· | ···················· |
| AA | 7029 | hCoV-19/Denmark/DCGC-18433/2020\|EPI_ISL_749965\|202 | ···················· | ···················· | ···················· | ···················· | ···················· | ···················· |
| AA | 7030 | hCoV-19/Denmark/DCGC-18438/2020\|EPI_ISL_749966\|202 | ···················· | ···················· | ···················· | ···················· | ···················· | ···················· |
| AA | 7031 | hCoV-19/Denmark/DCGC-18439/2020\|EPI_ISL_749967\|202 | ···················· | ···················· | ···················· | ···················· | ···················· | ···················· |
| AA | 7032 | hCoV-19/Denmark/DCGC-18442/2020\|EPI_ISL_749968\|202 | ···················· | ···················· | ···················· | ···················· | ···················· | ···················· |
| AA | 7033 | hCoV-19/Denmark/DCGC-18443/2020\|EPI_ISL_749969\|202 | ···················· | ···················· | ···················· | ···················· | ···················· | ···················· |
| AA | 7034 | hCoV-19/Denmark/DCGC-18444/2020\|EPI_ISL_749970\|202 | ···················· | ···················· | ···················· | ···················· | ···················· | ···················· |
| AA | 7035 | hCoV-19/Denmark/DCGC-18446/2020\|EPI_ISL_749971\|202 | ···················· | ···················· | ···················· | ···················· | ···················· | ···················· |
| AA | 7036 | hCoV-19/Denmark/DCGC-18447/2020\|EPI_ISL_749972\|202 | ···················· | ···················· | ···················· | ···················· | ···················· | ···················· |
| AA | 7037 | hCoV-19/Denmark/DCGC-18448/2020\|EPI_ISL_749973\|202 | ···················· | ···················· | ···················· | ···················· | ···················· | ···················· |
| AA | 7038 | hCoV-19/Denmark/DCGC-18449/2020\|EPI_ISL_749974\|202 | ···················· | ···················· | ···················· | ···················· | ···················· | ···················· |
| AA | 7039 | hCoV-19/Denmark/DCGC-18450/2020\|EPI_ISL_749975\|202 | ···················· | ···················· | ···················· | ···················· | ···················· | ···················· |
| AA | 7040 | hCoV-19/Denmark/DCGC-18451/2020\|EPI_ISL_749976\|202 | ···················· | ···················· | ···················· | ···················· | ···················· | ···················· |
| AA | 7041 | hCoV-19/Denmark/DCGC-18452/2020\|EPI_ISL_749977\|202 | ···················· | ···················· | ···················· | ···················· | ···················· | ···················· |
| AA | 7042 | hCoV-19/Denmark/DCGC-18453/2020\|EPI_ISL_749978\|202 | ···················· | ···················· | ···················· | ···················· | ···················· | ···················· |
| AA | 7043 | hCoV-19/Denmark/DCGC-18454/2020\|EPI_ISL_749979\|202 | ···················· | ···················· | ···················· | ···················· | ···················· | ···················· |
| AA | 7044 | hCoV-19/Denmark/DCGC-18458/2020\|EPI_ISL_749980\|202 | ···················· | ···················· | ···················· | ···················· | ···················· | ···················· |
| AA | 7045 | hCoV-19/Denmark/DCGC-18459/2020\|EPI_ISL_749981\|202 | ···················· | ···················· | ···················· | ···················· | ···················· | ···················· |
| AA | 7046 | hCoV-19/Denmark/DCGC-18462/2020\|EPI_ISL_749982\|202 | ···················· | ···················· | ···················· | ···················· | ···················· | ···················· |
| AA | 7047 | hCoV-19/Denmark/DCGC-18463/2020\|EPI_ISL_749983\|202 | ···················· | ···················· | ···················· | ···················· | ···················· | ···················· |
| AA | 7048 | hCoV-19/Denmark/DCGC-18466/2020\|EPI_ISL_749984\|202 | ···················· | ···················· | ···················· | ···················· | ···················· | ···················· |
| AA | 7049 | hCoV-19/Denmark/DCGC-18468/2020\|EPI_ISL_749985\|202 | ···················· | ···················· | ···················· | ···················· | ···················· | ···················· |
| AA | 7050 | hCoV-19/Denmark/DCGC-18472/2020\|EPI_ISL_749986\|202 | ···················· | ···················· | ···················· | ···················· | ···················· | ···················· |
| AA | 7051 | hCoV-19/Denmark/DCGC-18474/2020\|EPI_ISL_749987\|202 | ···················· | ·A·················· | ···················· | ···················· | ···················· | ··A················· |
| AA | 7052 | hCoV-19/Denmark/DCGC-18475/2020\|EPI_ISL_749988\|202 | ···················· | ·A·················· | ···················· | ···················· | ···················· | ··A················· |
| AA | 7053 | hCoV-19/Denmark/DCGC-18476/2020\|EPI_ISL_749989\|202 | ···················· | ···················· | ···················· | ···················· | ···················· | ···················· |
| AA | 7054 | hCoV-19/Denmark/DCGC-17327/2020\|EPI_ISL_750910\|202 | ···················· | ···················· | ···················· | ···················· | ···················· | ···················· |

| | | | | | | | | |
|---|---|---|---|---|---|---|---|---|
| AA | 7055 | hCoV-19/Denmark/DCGC-17329/2020\|EPI_ISL_750911\|202 | ···················· | ···················· | ···················· | ···················· | ···················· | ···················· |
| AA | 7056 | hCoV-19/Denmark/DCGC-17330/2020\|EPI_ISL_750912\|202 | ···················· | ···················· | ···················· | ···················· | ···················· | ···················· |
| AA | 7057 | hCoV-19/Denmark/DCGC-17331/2020\|EPI_ISL_750913\|202 | ···················· | ···················· | ···················· | ···················· | ···················· | ···················· |
| AA | 7058 | hCoV-19/Denmark/DCGC-17332/2020\|EPI_ISL_750914\|202 | ···················· | ···················· | ···················· | ···················· | ···················· | ···················· |
| AA | 7059 | hCoV-19/Denmark/DCGC-17333/2020\|EPI_ISL_750915\|202 | ···················· | ···················· | ···················· | ···················· | ···················· | ···················· |
| AA | 7060 | hCoV-19/Denmark/DCGC-17334/2020\|EPI_ISL_750916\|202 | ···················· | ···················· | ···················· | ···················· | ···················· | ···················· |
| AA | 7061 | hCoV-19/Denmark/DCGC-17335/2020\|EPI_ISL_750917\|202 | ···················· | ···················· | ···················· | ···················· | ···················· | ···················· |
| AA | 7062 | hCoV-19/Denmark/DCGC-17336/2020\|EPI_ISL_750918\|202 | ···················· | ···················· | ···················· | ···················· | ···················· | ···················· |
| AA | 7063 | hCoV-19/Denmark/DCGC-17337/2020\|EPI_ISL_750919\|202 | ···················· | ···················· | ···················· | ···················· | ···················· | ···················· |
| AA | 7064 | hCoV-19/Denmark/DCGC-17338/2020\|EPI_ISL_750920\|202 | ···················· | ···················· | ···················· | ···················· | ···················· | ···················· |
| AA | 7065 | hCoV-19/Denmark/DCGC-17340/2020\|EPI_ISL_750921\|202 | ···················· | ···················· | ···················· | ···················· | ···················· | ···················· |
| AA | 7066 | hCoV-19/Denmark/DCGC-17341/2020\|EPI_ISL_750922\|202 | ···················· | ···················· | ···················· | ···················· | ···················· | ···················· |
| AA | 7067 | hCoV-19/Denmark/DCGC-17343/2020\|EPI_ISL_750923\|202 | ···················· | ···················· | ···················· | ···················· | ···················· | ···················· |
| AA | 7068 | hCoV-19/Denmark/DCGC-17347/2020\|EPI_ISL_750924\|202 | ···················· | ···················· | ···················· | ···················· | ···················· | ···················· |
| AA | 7069 | hCoV-19/Denmark/DCGC-17348/2020\|EPI_ISL_750925\|202 | ···················· | ···················· | ···················· | ···················· | ···················· | ···················· |
| AA | 7070 | hCoV-19/Denmark/DCGC-17349/2020\|EPI_ISL_750926\|202 | ···················· | ···················· | ···················· | ···················· | ···················· | ···················· |
| AA | 7071 | hCoV-19/Denmark/DCGC-17352/2020\|EPI_ISL_750927\|202 | ···················· | ···················· | ···················· | ···················· | ···················· | ···················· |
| AA | 7072 | hCoV-19/Denmark/DCGC-17356/2020\|EPI_ISL_750928\|202 | ···················· | ···················· | ···················· | ···················· | ···················· | ···················· |
| AA | 7073 | hCoV-19/Denmark/DCGC-17357/2020\|EPI_ISL_750929\|202 | ···················· | ···················· | ···················· | ···················· | ···················· | ···················· |
| AA | 7074 | hCoV-19/Denmark/DCGC-17358/2020\|EPI_ISL_750930\|202 | ···················· | ···················· | ···················· | ···················· | ···················· | ···················· |
| AA | 7075 | hCoV-19/Denmark/DCGC-17359/2020\|EPI_ISL_750931\|202 | ···················· | ···················· | ···················· | ···················· | ···················· | ···················· |
| AA | 7076 | hCoV-19/Denmark/DCGC-17360/2020\|EPI_ISL_750932\|202 | ···················· | ···················· | ···················· | ···················· | ···················· | ···················· |
| AA | 7077 | hCoV-19/Denmark/DCGC-17361/2020\|EPI_ISL_750933\|202 | ···················· | ···················· | ···················· | ···················· | ···················· | ···················· |
| AA | 7078 | hCoV-19/Denmark/DCGC-17363/2020\|EPI_ISL_750934\|202 | ···················· | ···················· | ···················· | ···················· | ···················· | ···················· |
| AA | 7079 | hCoV-19/Denmark/DCGC-17365/2020\|EPI_ISL_750935\|202 | ···················· | ···················· | ···················· | ···················· | ···················· | ···················· |
| AA | 7080 | hCoV-19/Denmark/DCGC-17366/2020\|EPI_ISL_750936\|202 | ···················· | ···················· | ···················· | ···················· | ···················· | ···················· |
| AA | 7081 | hCoV-19/Denmark/DCGC-17367/2020\|EPI_ISL_750937\|202 | ···················· | ···················· | ···················· | ···················· | ···················· | ···················· |
| AA | 7082 | hCoV-19/Denmark/DCGC-17369/2020\|EPI_ISL_750938\|202 | ···················· | ···················· | ···················· | ···················· | ···················· | ···················· |
| AA | 7083 | hCoV-19/Denmark/DCGC-17371/2020\|EPI_ISL_750939\|202 | ···················· | ·A·················· | ···················· | ···················· | ···················· | ··A················· |
| AA | 7084 | hCoV-19/USA/HI-H200657/2020\|EPI_ISL_753180\|2020-12 | ···················· | ·A·················· | ···················· | ···················· | ···················· | ···················· |
| AA | 7085 | hCoV-19/USA/HI-H200658/2020\|EPI_ISL_753181\|2020-12 | ···················· | ·A·················· | ···················· | ···················· | ···················· | ···················· |
| AA | 7086 | hCoV-19/USA/HI-H200659/2020\|EPI_ISL_753182\|2020-12 | ···················· | ·A·················· | ···················· | ···················· | ···················· | ···················· |
| AA | 7087 | hCoV-19/USA/HI-H200660/2020\|EPI_ISL_753183\|2020-12 | ···················· | ·A·················· | ···················· | ···················· | ···················· | ···················· |
| AA | 7088 | hCoV-19/USA/HI-H200661/2020\|EPI_ISL_753184\|2020-12 | ···················· | ·A·················· | ···················· | ···················· | ···················· | ···················· |
| AA | 7089 | hCoV-19/USA/HI-H200662/2020\|EPI_ISL_753185\|2020-12 | ···················· | ·A·················· | ···················· | ···················· | ···················· | ···················· |
| AA | 7090 | hCoV-19/USA/HI-H200663/2020\|EPI_ISL_753186\|2020-12 | ···················· | ·A·················· | ···················· | ···················· | ···················· | ···················· |
| AA | 7091 | hCoV-19/USA/HI-H200664/2020\|EPI_ISL_753187\|2020-12 | ···················· | ·A·················· | ···················· | ···················· | ···················· | ···················· |
| AA | 7092 | hCoV-19/USA/HI-H200665/2020\|EPI_ISL_753188\|2020-12 | ···················· | ·A·················· | ···················· | ···················· | ···················· | ···················· |
| AA | 7093 | hCoV-19/USA/HI-H200666/2020\|EPI_ISL_753189\|2020-12 | ···················· | ·A·················· | ···················· | ···················· | ···················· | ···················· |
| AA | 7094 | hCoV-19/USA/HI-H200667/2020\|EPI_ISL_753190\|2020-12 | ···················· | ·A·················· | ···················· | ···················· | ···················· | ···················· |
| AA | 7095 | hCoV-19/USA/HI-H200668/2020\|EPI_ISL_753191\|2020-12 | ···················· | ·A·················· | ···················· | ···················· | ···················· | ···················· |
| AA | 7096 | hCoV-19/USA/HI-H200669/2020\|EPI_ISL_753192\|2020-09 | ···················· | ·A·················· | ···················· | ···················· | ···················· | ···················· |
| AA | 7097 | hCoV-19/USA/HI-H200670/2020\|EPI_ISL_753193\|2020-09 | ···················· | ·A·················· | ···················· | ···················· | ···················· | ···················· |
| AA | 7098 | hCoV-19/USA/HI-H200671/2020\|EPI_ISL_753194\|2020-09 | ···················· | ·A·················· | ···················· | ···················· | ···················· | ···················· |
| AA | 7099 | hCoV-19/USA/HI-H200672/2020\|EPI_ISL_753195\|2020-09 | ···················· | ·A·················· | ···················· | ···················· | ···················· | ···················· |
| AA | 7100 | hCoV-19/USA/HI-H200673/2020\|EPI_ISL_753196\|2020-11 | ···················· | ·A·················· | ···················· | ···················· | ···················· | ···················· |
| AA | 7101 | hCoV-19/USA/HI-H200674/2020\|EPI_ISL_753197\|2020-11 | ···················· | ·A·················· | ···················· | ···················· | ···················· | ···················· |
| AA | 7102 | hCoV-19/USA/HI-H200675/2020\|EPI_ISL_753198\|2020-11 | ···················· | ·A·················· | ···················· | ···················· | ···················· | ···················· |
| AA | 7103 | hCoV-19/USA/HI-H200676/2020\|EPI_ISL_753199\|2020-11 | ···················· | ·A·················· | ···················· | ···················· | ···················· | ···················· |
| AA | 7104 | hCoV-19/Denmark/DCGC-22028/2020\|EPI_ISL_758030\|202 | ···················· | ···················· | ···················· | ···················· | ··C················· | ···················· |
| AA | 7105 | hCoV-19/Denmark/DCGC-22044/2020\|EPI_ISL_758031\|202 | ···················· | ···················· | ···················· | ···················· | ···················· | ···················· |
| AA | 7106 | hCoV-19/Denmark/DCGC-22048/2020\|EPI_ISL_758032\|202 | ···················· | ···················· | ···················· | ···················· | ···················· | ···················· |
| AA | 7107 | hCoV-19/Denmark/DCGC-22060/2020\|EPI_ISL_758033\|202 | ···················· | ···················· | ···················· | ···················· | ···················· | ···················· |
| AA | 7108 | hCoV-19/Denmark/DCGC-22065/2020\|EPI_ISL_758034\|202 | ···················· | ···················· | ···················· | ···················· | ···················· | ···················· |
| AA | 7109 | hCoV-19/Denmark/DCGC-22067/2020\|EPI_ISL_758035\|202 | ···················· | ···················· | ···················· | ···················· | ···················· | ···················· |
| AA | 7110 | hCoV-19/Denmark/DCGC-22072/2020\|EPI_ISL_758036\|202 | ···················· | ···················· | ···················· | ···················· | ···················· | ···················· |
| AA | 7111 | hCoV-19/Denmark/DCGC-22073/2020\|EPI_ISL_758037\|202 | ···················· | ···················· | ···················· | ···················· | ···················· | ···················· |
| AA | 7112 | hCoV-19/Denmark/DCGC-22095/2020\|EPI_ISL_758038\|202 | ···················· | ···················· | ···················· | ···················· | ···················· | ···················· |
| AA | 7113 | hCoV-19/Denmark/DCGC-22097/2020\|EPI_ISL_758039\|202 | ···················· | ···················· | ···················· | ···················· | ···················· | ···················· |
| AA | 7114 | hCoV-19/Denmark/DCGC-22104/2020\|EPI_ISL_758040\|202 | ···················· | ···················· | ···················· | ···················· | ···················· | ···················· |
| AA | 7115 | hCoV-19/Denmark/DCGC-22106/2020\|EPI_ISL_758041\|202 | ···················· | ···················· | ···················· | ···················· | ···················· | ···················· |
| AA | 7116 | hCoV-19/Denmark/DCGC-22122/2020\|EPI_ISL_758042\|202 | ···················· | ···················· | ···················· | ···················· | ···················· | ···················· |
| AA | 7117 | hCoV-19/Denmark/DCGC-22142/2020\|EPI_ISL_758043\|202 | ···················· | ···················· | ···················· | ···················· | ···················· | ···················· |
| AA | 7118 | hCoV-19/Denmark/DCGC-22151/2020\|EPI_ISL_758044\|202 | ···················· | ··G················· | ···················· | ···················· | ···················· | ···················· |
| AA | 7119 | hCoV-19/Denmark/DCGC-22155/2020\|EPI_ISL_758045\|202 | ···················· | ···················· | ···················· | ···················· | ···················· | ···················· |
| AA | 7120 | hCoV-19/Denmark/DCGC-22159/2020\|EPI_ISL_758046\|202 | ···················· | ···················· | ···················· | ···················· | ···················· | ···················· |
| AA | 7121 | hCoV-19/Denmark/DCGC-22165/2020\|EPI_ISL_758047\|202 | ···················· | ···················· | ···················· | ···················· | ···················· | ···················· |
| AA | 7122 | hCoV-19/Denmark/DCGC-22167/2020\|EPI_ISL_758048\|202 | ···················· | ···················· | ···················· | ···················· | ···················· | ···················· |
| AA | 7123 | hCoV-19/Denmark/DCGC-22192/2020\|EPI_ISL_758049\|202 | ···················· | ···················· | ···················· | ···················· | ···················· | ···················· |
| AA | 7124 | hCoV-19/Denmark/DCGC-22200/2020\|EPI_ISL_758050\|202 | ···················· | ···················· | ···················· | ···················· | ···················· | ···················· |
| AA | 7125 | hCoV-19/Denmark/DCGC-22206/2020\|EPI_ISL_758051\|202 | ···················· | ···················· | ···················· | ···················· | ···················· | ···················· |
| AA | 7126 | hCoV-19/Denmark/DCGC-22228/2020\|EPI_ISL_758052\|202 | ···················· | ··G················· | ···················· | ···················· | ···················· | ···················· |
| AA | 7127 | hCoV-19/Denmark/DCGC-22231/2020\|EPI_ISL_758053\|202 | ···················· | ···················· | ···················· | ···················· | ···················· | ···················· |
| AA | 7128 | hCoV-19/Denmark/DCGC-22237/2020\|EPI_ISL_758054\|202 | ···················· | ···················· | ···················· | ···················· | ···················· | ···················· |
| AA | 7129 | hCoV-19/Denmark/DCGC-22238/2020\|EPI_ISL_758055\|202 | ···················· | ···················· | ···················· | ···················· | ···················· | ···················· |
| AA | 7130 | hCoV-19/Denmark/DCGC-22240/2020\|EPI_ISL_758056\|202 | ···················· | ···················· | ···················· | ···················· | ···················· | ···················· |
| AA | 7131 | hCoV-19/Denmark/DCGC-22242/2020\|EPI_ISL_758057\|202 | ···················· | ···················· | ···················· | ···················· | ···················· | ···················· |
| AA | 7132 | hCoV-19/Denmark/DCGC-22250/2020\|EPI_ISL_758058\|202 | ···················· | ···················· | ···················· | ···················· | ···················· | ···················· |
| AA | 7133 | hCoV-19/Denmark/DCGC-22271/2020\|EPI_ISL_758059\|202 | ···················· | ···················· | ···················· | ···················· | ···················· | ···················· |
| AA | 7134 | hCoV-19/Denmark/DCGC-22276/2020\|EPI_ISL_758060\|202 | ···················· | ···················· | ···················· | ···················· | ···················· | ···················· |

| | | | | | | | | |
|---|---|---|---|---|---|---|---|---|
| AA | 7135 | hCoV-19/Denmark/DCGC-22284/2020\|EPI_ISL_758061\|202 | ···················· | ···················· | ····················· | ···················· | ···C················ | ···················· |
| AA | 7136 | hCoV-19/Denmark/DCGC-20783/2020\|EPI_ISL_759360\|202 | ···················· | ···················· | ····················· | ···················· | ···················· | ···················· |
| AA | 7137 | hCoV-19/Denmark/DCGC-20785/2020\|EPI_ISL_759361\|202 | ···················· | ···················· | ····················· | ···················· | ···················· | ···················· |
| AA | 7138 | hCoV-19/Denmark/DCGC-20786/2020\|EPI_ISL_759362\|202 | ···················· | ···················· | ····················· | ···················· | ···················· | ···················· |
| AA | 7139 | hCoV-19/Denmark/DCGC-20787/2020\|EPI_ISL_759363\|202 | ···················· | ···················· | ····················· | ···················· | ···················· | ···················· |
| AA | 7140 | hCoV-19/Denmark/DCGC-20789/2020\|EPI_ISL_759364\|202 | ···················· | ···················· | ····················· | ···················· | ···················· | ···················· |
| AA | 7141 | hCoV-19/Denmark/DCGC-20790/2020\|EPI_ISL_759365\|202 | ···················· | ···················· | ····················· | ···················· | ···················· | ···················· |
| AA | 7142 | hCoV-19/Denmark/DCGC-20791/2020\|EPI_ISL_759366\|202 | ···················· | ···················· | ····················· | ···················· | ···················· | ···················· |
| AA | 7143 | hCoV-19/Denmark/DCGC-20792/2020\|EPI_ISL_759367\|202 | ···················· | ···················· | ····················· | ···················· | ···················· | ···················· |
| AA | 7144 | hCoV-19/Denmark/DCGC-20793/2020\|EPI_ISL_759368\|202 | ···················· | ···················· | ····················· | ···················· | ···················· | ···················· |
| AA | 7145 | hCoV-19/Denmark/DCGC-20797/2020\|EPI_ISL_759369\|202 | ···················· | ···················· | ····················· | ···················· | ···················· | ···················· |
| AA | 7146 | hCoV-19/Denmark/DCGC-20799/2020\|EPI_ISL_759370\|202 | ···················· | ···················· | ····················· | ···················· | ···················· | ···················· |
| AA | 7147 | hCoV-19/Denmark/DCGC-20800/2020\|EPI_ISL_759371\|202 | ·······C············ | ···················· | ····················· | ···················· | ···················· | ···················· |
| AA | 7148 | hCoV-19/Denmark/DCGC-20801/2020\|EPI_ISL_759372\|202 | ···················· | ···················· | ····················· | ···················· | ···················· | ···················· |
| AA | 7149 | hCoV-19/Denmark/DCGC-20804/2020\|EPI_ISL_759373\|202 | ···················· | ···················· | ····················· | ···················· | ···················· | ···················· |
| AA | 7150 | hCoV-19/Denmark/DCGC-20805/2020\|EPI_ISL_759374\|202 | ···················· | ···················· | ····················· | ···················· | ···················· | ···················· |
| AA | 7151 | hCoV-19/Denmark/DCGC-20807/2020\|EPI_ISL_759375\|202 | ···················· | ···················· | ····················· | ···················· | ···················· | ···················· |
| AA | 7152 | hCoV-19/Denmark/DCGC-20808/2020\|EPI_ISL_759376\|202 | ···················· | ···················· | ····················· | ···················· | ···················· | ···················· |
| AA | 7153 | hCoV-19/Denmark/DCGC-20809/2020\|EPI_ISL_759377\|202 | ···················· | ···················· | ····················· | ···················· | ···················· | ···················· |
| AA | 7154 | hCoV-19/Denmark/DCGC-20810/2020\|EPI_ISL_759378\|202 | ···················· | ·A·················· | ····················· | ···················· | ···················· | ···················· |
| AA | 7155 | hCoV-19/Denmark/DCGC-20812/2020\|EPI_ISL_759379\|202 | ···················· | ···················· | ····················· | ···················· | ···················· | ···················· |
| AA | 7156 | hCoV-19/Denmark/DCGC-20813/2020\|EPI_ISL_759380\|202 | ···················· | ··· A··············· | ····················· | ···················· | ···················· | ···················· |
| AA | 7157 | hCoV-19/Denmark/DCGC-20814/2020\|EPI_ISL_759381\|202 | ···················· | ···················· | ····················· | ···················· | ···················· | ···················· |
| AA | 7158 | hCoV-19/Denmark/DCGC-20817/2020\|EPI_ISL_759382\|202 | ···················· | ·A·················· | ····················· | ···················· | ···················· | ···A················ |
| AA | 7159 | hCoV-19/Denmark/DCGC-20818/2020\|EPI_ISL_759383\|202 | ···················· | ···················· | ····················· | ···················· | ···················· | ···················· |
| AA | 7160 | hCoV-19/Denmark/DCGC-20819/2020\|EPI_ISL_759384\|202 | ···················· | ···················· | ····················· | ···················· | ···················· | ···················· |
| AA | 7161 | hCoV-19/Denmark/DCGC-20822/2020\|EPI_ISL_759385\|202 | ···················· | ···················· | ····················· | ···················· | ···················· | ···················· |
| AA | 7162 | hCoV-19/Denmark/DCGC-20823/2020\|EPI_ISL_759386\|202 | ···················· | ···················· | ····················· | ···················· | ···················· | ···················· |
| AA | 7163 | hCoV-19/Denmark/DCGC-20824/2020\|EPI_ISL_759387\|202 | ···················· | ···················· | ····················· | ···················· | ···················· | ···················· |
| AA | 7164 | hCoV-19/Denmark/DCGC-20825/2020\|EPI_ISL_759388\|202 | ···················· | ···················· | ····················· | ···················· | ···················· | ···················· |
| AA | 7165 | hCoV-19/Denmark/DCGC-20828/2020\|EPI_ISL_759389\|202 | ···················· | ···················· | ····················· | ···················· | ···················· | ···················· |
| AA | 7166 | hCoV-19/Denmark/DCGC-20830/2020\|EPI_ISL_759390\|202 | ···················· | ···················· | ····················· | ···················· | ···················· | ···················· |
| AA | 7167 | hCoV-19/Denmark/DCGC-20832/2020\|EPI_ISL_759391\|202 | ···················· | ···················· | ····················· | ···················· | ···················· | ···················· |
| AA | 7168 | hCoV-19/Denmark/DCGC-20833/2020\|EPI_ISL_759392\|202 | ···················· | ···················· | ····················· | ···················· | ···················· | ···················· |
| AA | 7169 | hCoV-19/South Korea/KDCA0104/2020\|EPI_ISL_747340\|2 | ···················· | ·A·················· | ····················· | ···················· | ···················· | ···A················ |
| AA | 7170 | hCoV-19/South Korea/KDCA0105/2020\|EPI_ISL_747341\|2 | ···················· | ···················· | ····················· | ···················· | ···················· | ···················· |
| AA | 7171 | hCoV-19/South Korea/KDCA0107/2020\|EPI_ISL_747343\|2 | ···················· | ···················· | ····················· | ···················· | ···················· | ···A················ |
| AA | 7172 | hCoV-19/South Korea/KDCA0108/2020\|EPI_ISL_747344\|2 | ···················· | ···················· | ····················· | ···················· | ···················· | ···················· |
| AA | 7173 | hCoV-19/South Korea/KDCA0114/2020\|EPI_ISL_747349\|2 | ···················· | ···················· | ····················· | ···················· | ···················· | ···················· |
| AA | 7174 | hCoV-19/South Korea/KDCA0145/2020\|EPI_ISL_747380\|2 | ···················· | ···················· | ····················· | ···················· | ···················· | ···················· |
| AA | 7175 | hCoV-19/South Korea/KDCA0146/2020\|EPI_ISL_747381\|2 | ···················· | ···················· | ····················· | ···················· | ···················· | ···················· |
| AA | 7176 | hCoV-19/South Korea/KDCA0147/2020\|EPI_ISL_747382\|2 | ···················· | ···················· | ····················· | ···················· | ···················· | ···················· |
| AA | 7178 | hCoV-19/South Korea/KDCA0150/2020\|EPI_ISL_747384\|2 | ···················· | ···················· | ····················· | ···················· | ···················· | ···················· |
| AA | 7179 | hCoV-19/South Korea/KDCA0151/2020\|EPI_ISL_747385\|2 | ···················· | ···················· | ····················· | ···················· | ···················· | ···················· |
| AA | 7180 | hCoV-19/South Korea/KDCA0152/2020\|EPI_ISL_747386\|2 | ···················· | ···················· | ····················· | ···················· | ···················· | ···················· |
| AA | 7181 | hCoV-19/South Korea/KDCA0153/2020\|EPI_ISL_747387\|2 | ···················· | ···················· | ····················· | ···················· | ···················· | ···················· |
| AA | 7182 | hCoV-19/South Korea/KDCA0154/2020\|EPI_ISL_747388\|2 | ···················· | ···················· | ····················· | ···················· | ···················· | ···················· |
| AA | 7183 | hCoV-19/South Korea/KDCA0155/2020\|EPI_ISL_747389\|2 | ···················· | ···················· | ····················· | ···················· | ···················· | ···················· |
| AA | 7184 | hCoV-19/South Korea/KDCA0156/2020\|EPI_ISL_747390\|2 | ···················· | ···················· | ····················· | ···················· | ···················· | ···················· |
| AA | 7185 | hCoV-19/South Korea/KDCA0157/2020\|EPI_ISL_747391\|2 | ···················· | ···················· | ····················· | ···················· | ···················· | ···················· |
| AA | 7186 | hCoV-19/South Korea/KDCA0159/2020\|EPI_ISL_747393\|2 | ···················· | ···················· | ····················· | ···················· | ···················· | ···················· |
| AA | 7187 | hCoV-19/South Korea/KDCA0160/2020\|EPI_ISL_747394\|2 | ···················· | ···················· | ····················· | ···················· | ···················· | ···················· |
| AA | 7188 | hCoV-19/South Korea/KDCA0161/2020\|EPI_ISL_747395\|2 | ···················· | ···················· | ····················· | ···················· | ···················· | ···················· |
| AA | 7189 | hCoV-19/South Korea/KDCA0162/2020\|EPI_ISL_747396\|2 | ···················· | ···················· | ····················· | ···················· | ···················· | ···················· |
| AA | 7190 | hCoV-19/South Korea/KDCA0163/2020\|EPI_ISL_747397\|2 | ···················· | ···················· | ····················· | ···················· | ···················· | ···················· |
| AA | 7191 | hCoV-19/South Korea/KDCA0164/2020\|EPI_ISL_747398\|2 | ···················· | ···················· | ····················· | ···················· | ···················· | ···················· |
| AA | 7192 | hCoV-19/South Korea/KDCA0165/2020\|EPI_ISL_747399\|2 | ···················· | ···················· | ····················· | ···················· | ···················· | ···················· |
| AA | 7193 | hCoV-19/Denmark/DCGC-18965/2020\|EPI_ISL_748670\|202 | ···················· | ···················· | ····················· | ···················· | ···················· | ···················· |
| AA | 7194 | hCoV-19/Denmark/DCGC-18967/2020\|EPI_ISL_748671\|202 | ···················· | ·A·················· | ····················· | ···················· | ···················· | ···A················ |
| AA | 7195 | hCoV-19/Denmark/DCGC-18970/2020\|EPI_ISL_748672\|202 | ···················· | ···················· | ····················· | ···················· | ···················· | ···················· |
| AA | 7196 | hCoV-19/Denmark/DCGC-18971/2020\|EPI_ISL_748673\|202 | ···················· | ···················· | ····················· | ···················· | ···················· | ···················· |
| AA | 7197 | hCoV-19/Denmark/DCGC-18972/2020\|EPI_ISL_748674\|202 | ···················· | ···················· | ····················· | ···················· | ···················· | ···················· |
| AA | 7198 | hCoV-19/Denmark/DCGC-18973/2020\|EPI_ISL_748675\|202 | ···················· | ···················· | ····················· | ···················· | ···················· | ···················· |
| AA | 7199 | hCoV-19/Denmark/DCGC-17439/2020\|EPI_ISL_750983\|202 | ···················· | ···················· | ····················· | ···················· | ···················· | ···················· |
| AA | 7200 | hCoV-19/Denmark/DCGC-17440/2020\|EPI_ISL_750984\|202 | ···················· | ···················· | ····················· | ···················· | ···················· | ···················· |
| AA | 7201 | hCoV-19/Denmark/DCGC-17441/2020\|EPI_ISL_750985\|202 | ···················· | ···················· | ····················· | ···················· | ···················· | ···················· |
| AA | 7202 | hCoV-19/Denmark/DCGC-17443/2020\|EPI_ISL_750986\|202 | ···················· | ···················· | ····················· | ···················· | ···················· | ···················· |
| AA | 7203 | hCoV-19/Denmark/DCGC-17444/2020\|EPI_ISL_750987\|202 | ···················· | ···················· | ····················· | ···················· | ···················· | ···················· |
| AA | 7204 | hCoV-19/Denmark/DCGC-17445/2020\|EPI_ISL_750988\|202 | ···················· | ···················· | ····················· | ···················· | ···················· | ···················· |
| AA | 7205 | hCoV-19/Denmark/DCGC-17446/2020\|EPI_ISL_750989\|202 | ···················· | ···················· | ····················· | ···················· | ···················· | ···················· |
| AA | 7206 | hCoV-19/Denmark/DCGC-17447/2020\|EPI_ISL_750990\|202 | ···················· | ···················· | ····················· | ···················· | ···················· | ···················· |
| AA | 7207 | hCoV-19/Denmark/DCGC-17448/2020\|EPI_ISL_750991\|202 | ···················· | ···················· | ····················· | ···················· | ···················· | ···················· |
| AA | 7208 | hCoV-19/Denmark/DCGC-17449/2020\|EPI_ISL_750992\|202 | ···················· | ···················· | ····················· | ···················· | ···················· | ···················· |
| AA | 7209 | hCoV-19/Denmark/DCGC-17452/2020\|EPI_ISL_750993\|202 | ···················· | ·A·················· | ····················· | ···················· | ···················· | ···A················ |
| AA | 7210 | hCoV-19/Denmark/DCGC-17454/2020\|EPI_ISL_750994\|202 | ···················· | ···················· | ····················· | ···················· | ···················· | ···················· |
| AA | 7211 | hCoV-19/Denmark/DCGC-17456/2020\|EPI_ISL_750995\|202 | ···················· | ···················· | ····················· | ···················· | ···················· | ···················· |
| AA | 7212 | hCoV-19/Denmark/DCGC-17457/2020\|EPI_ISL_750996\|202 | ···················· | ···················· | ····················· | ···················· | ···················· | ···················· |
| AA | 7213 | hCoV-19/Denmark/DCGC-17459/2020\|EPI_ISL_750997\|202 | ···················· | ···················· | ····················· | ···················· | ···················· | ···················· |
| AA | 7214 | hCoV-19/Denmark/DCGC-17462/2020\|EPI_ISL_750998\|202 | ···················· | ···················· | ····················· | ···················· | ···················· | ···················· |
| AA | 7215 | hCoV-19/Denmark/DCGC-17464/2020\|EPI_ISL_750999\|202 | ···················· | ···················· | ····················· | ···················· | ···················· | ···················· |

AA 7216 hCoV-19/USA/HI-H200677/2020|EPI_ISL_753200|2020-11 ···················· ···················· ···················· ···················· ···················· ····················
AA 7217 hCoV-19/USA/HI-H200678/2020|EPI_ISL_753201|2020-11 ···················· ···················· ···················· ···················· ···················· ····················
AA 7218 hCoV-19/USA/HI-H200679/2020|EPI_ISL_753202|2020-12 ···················· ·A·················· ···················· ···················· ···················· ····················
AA 7219 hCoV-19/USA/HI-H200680/2020|EPI_ISL_753203|2020-12 ···················· ·A·················· ···················· ···················· ···················· ····················
AA 7220 hCoV-19/USA/HI-H200681/2020|EPI_ISL_753204|2020-12 ···················· ·A·················· ···················· ···················· ···················· ····················
AA 7221 hCoV-19/USA/HI-H200682/2020|EPI_ISL_753205|2020-12 ···················· ·A·················· ···················· ···················· ···················· ····················
AA 7222 hCoV-19/USA/HI-H200683/2020|EPI_ISL_753206|2020-12 ···················· ·A·················· ···················· ···················· ···················· ····················
AA 7223 hCoV-19/USA/HI-H200684/2020|EPI_ISL_753207|2020-12 ···················· ·A·················· ···················· ···················· ···················· ····················
AA 7224 hCoV-19/USA/HI-H200685/2020|EPI_ISL_753208|2020-12 ···················· ·A·················· ···················· ···················· ···················· ····················
AA 7225 hCoV-19/USA/HI-H200686/2020|EPI_ISL_753209|2020-12 ···················· ·A·················· ···················· ···················· ···················· ····················
AA 7226 hCoV-19/USA/HI-H200687/2020|EPI_ISL_753210|2020-12 ···················· ·A·················· ···················· ···················· ···················· ····················
AA 7227 hCoV-19/USA/HI-H200688/2020|EPI_ISL_753211|2020-12 ···················· ·A·················· ···················· ···················· ···················· ····················
AA 7228 hCoV-19/USA/HI-H200690/2020|EPI_ISL_753212|2020-12 ···················· ·A·················· ···················· ···················· ···················· ····················
AA 7229 hCoV-19/USA/HI-H200691/2020|EPI_ISL_753213|2020-12 ···················· ·A·················· ···················· ···················· ···················· ····················
AA 7230 hCoV-19/USA/HI-H200692/2020|EPI_ISL_753214|2020-11 ···················· ···················· ···················· ···················· ···················· ····················
AA 7231 hCoV-19/USA/HI-H200693/2020|EPI_ISL_753215|2020-09 ···················· ·A·················· ···················· ···················· ···················· ····················
AA 7232 hCoV-19/USA/HI-H200739/2020|EPI_ISL_753260|2020-12 ···················· ·A·················· ···················· ···················· ···················· ····················
AA 7233 hCoV-19/USA/HI-H200740/2020|EPI_ISL_753261|2020-12 ···················· ·A·················· ···················· ···················· ···················· ····················
AA 7234 hCoV-19/USA/CA-CHLA-PLM30832286/2020|EPI_ISL_75326 ···················· ···················· ···················· ···················· ···················· ····················
AA 7235 hCoV-19/USA/CA-CHLA-PLM06076230/2020|EPI_ISL_75326 ···················· ·A·················· ···················· ···················· ···················· ····················
AA 7236 hCoV-19/USA/CA-CHLA-PLM66641292/2020|EPI_ISL_75326 ···················· ···················· ···················· ···················· ···················· ····················
AA 7237 hCoV-19/USA/CA-CHLA-PLM33142446/2020|EPI_ISL_75326 ···················· ·A·················· ···················· ···················· ···················· ····················
AA 7238 hCoV-19/USA/CA-CHLA-PLM89994435/2020|EPI_ISL_75326 ···················· ·A·················· ···················· ···················· ···················· ····················
AA 7239 hCoV-19/USA/CA-CHLA-PLM49702170/2020|EPI_ISL_75326 ···················· ·A·················· ···················· ···················· ···················· ····················
AA 7240 hCoV-19/USA/CA-CHLA-PLM31468132/2020|EPI_ISL_75326 ···················· ·A·················· ···················· ···················· ···················· ····················
AA 7241 hCoV-19/USA/CA-CHLA-PLM78795231/2020|EPI_ISL_75327 ···················· ·A·················· ···················· ···················· ···················· ····················
AA 7242 hCoV-19/USA/CA-CHLA-PLM15535800/2020|EPI_ISL_75327 ···················· ·A·················· ···················· ···················· ···················· ····················
AA 7243 hCoV-19/USA/CA-CHLA-PLM26509492/2020|EPI_ISL_75327 ···················· ···················· ···················· ···················· ···················· ····················
AA 7244 hCoV-19/USA/CA-CHLA-PLM00400834/2020|EPI_ISL_75327 ···················· ·A·················· ···················· ···················· ···················· ····················
AA 7245 hCoV-19/USA/NY-Wadsworth-280462-01/2020|EPI_ISL_75 ···················· ···················· ···················· ···················· ···················· ····················
AA 7246 hCoV-19/USA/NY-Wadsworth-280441-01/2020|EPI_ISL_75 ···················· ···················· ···················· ···················· ···················· ····················
AA 7247 hCoV-19/USA/NY-Wadsworth-280476-01/2020|EPI_ISL_75 ···················· ···················· ···················· ···················· ···················· ····················
AA 7248 hCoV-19/USA/NY-Wadsworth-280464-01/2020|EPI_ISL_75 ···················· ···················· ···················· ···················· ···················· ····················
AA 7249 hCoV-19/USA/NY-Wadsworth-281268-01/2020|EPI_ISL_75 ···················· ···················· ···················· ···················· ···················· ····················
AA 7250 hCoV-19/USA/NY-Wadsworth-281330-01/2020|EPI_ISL_75 ···················· ·A·················· ···················· ···················· ···················· ····················
AA 7251 hCoV-19/USA/NY-Wadsworth-280805-01/2020|EPI_ISL_75 ···················· ·A·················· ···················· ···················· ···················· ····················
AA 7252 hCoV-19/USA/NY-Wadsworth-280806-01/2020|EPI_ISL_75 ···················· ·A·················· ···················· ···················· ···················· ····················
AA 7253 hCoV-19/USA/NY-Wadsworth-280814-01/2020|EPI_ISL_75 ···················· ···················· ···················· ···················· ···················· ····················
AA 7254 hCoV-19/USA/NY-Wadsworth-280797-01/2020|EPI_ISL_75 ···················· ···················· ···················· ···················· ···················· ····················
AA 7255 hCoV-19/USA/NY-Wadsworth-261032-01/2020|EPI_ISL_75 ···················· ···················· ···················· ···················· ···················· ····················
AA 7256 hCoV-19/USA/NY-Wadsworth-261037-01/2020|EPI_ISL_75 ···················· ···················· ···················· ···················· ···················· ····················
AA 7257 hCoV-19/USA/NY-Wadsworth-261035-01/2020|EPI_ISL_75 ···················· ···················· ···················· ···················· ···················· ····················
AA 7258 hCoV-19/USA/NY-Wadsworth-263735-01/2020|EPI_ISL_75 ···················· ·A·················· ···················· ···················· ···················· ····················
AA 7259 hCoV-19/USA/NY-Wadsworth-263748-01/2020|EPI_ISL_75 ···················· ·············A······ ···················· ···················· ···················· ····················
AA 7260 hCoV-19/USA/NY-Wadsworth-263950-01/2020|EPI_ISL_75 ···················· ···················· ···················· ···················· ···················· ····················
AA 7261 hCoV-19/USA/NY-Wadsworth-263927-01/2020|EPI_ISL_75 ···················· ·A·················· ···················· ···················· ···················· ····················
AA 7262 hCoV-19/USA/NY-Wadsworth-264206-01/2020|EPI_ISL_75 ···················· ·A·················· ···················· ···················· ···················· ····················
AA 7263 hCoV-19/USA/NY-Wadsworth-264207-01/2020|EPI_ISL_75 ···················· ·A·················· ···················· ···················· ···················· ····················
AA 7264 hCoV-19/USA/NY-Wadsworth-264208-01/2020|EPI_ISL_75 ···················· ·A·················· ···················· ···················· ···················· ····················
AA 7265 hCoV-19/USA/NY-Wadsworth-264211-01/2020|EPI_ISL_75 ···················· ·A·················· ···················· ···················· ···················· ····················
AA 7266 hCoV-19/USA/NY-Wadsworth-264196-01/2020|EPI_ISL_75 ···················· ·A·················· ···················· ···················· ···················· ····················
AA 7267 hCoV-19/USA/NY-Wadsworth-264200-01/2020|EPI_ISL_75 ···················· ·A·················· ···················· ···················· ···················· ····················
AA 7268 hCoV-19/USA/NY-Wadsworth-264201-01/2020|EPI_ISL_75 ···················· ·A·················· ···················· ···················· ···················· ····················
AA 7269 hCoV-19/USA/NY-Wadsworth-264290-01/2020|EPI_ISL_75 ···················· ·A·················· ···················· ···················· ···················· ····················
AA 7270 hCoV-19/USA/NY-Wadsworth-268995-01/2020|EPI_ISL_75 ···················· ·A·················· ···················· ···················· ···················· ····················
AA 7271 hCoV-19/USA/NY-Wadsworth-233786-01/2020|EPI_ISL_75 ···················· ·············A······ ···················· ···················· ···················· ····················
AA 7272 hCoV-19/USA/NY-Wadsworth-233496-01/2020|EPI_ISL_75 ···················· ···················· ···················· ···················· ···················· ····················
AA 7273 hCoV-19/USA/NY-Wadsworth-233510-01/2020|EPI_ISL_75 ···················· ·A·················· ···················· ···················· ···················· ····················
AA 7274 hCoV-19/USA/NY-Wadsworth-233298-01/2020|EPI_ISL_75 ···················· ·A·················· ···················· ···················· ···················· ····················
AA 7275 hCoV-19/USA/NY-Wadsworth-233903-01/2020|EPI_ISL_75 ···················· ···················· ···················· ···················· ···················· ····················
AA 7276 hCoV-19/USA/NY-Wadsworth-233889-01/2020|EPI_ISL_75 ···················· ···················· ···················· ···················· ···················· ····················
AA 7277 hCoV-19/USA/NY-Wadsworth-233890-01/2020|EPI_ISL_75 ···················· ···················· ···················· ···················· ···················· ····················
AA 7278 hCoV-19/USA/NY-Wadsworth-236445-01/2020|EPI_ISL_75 ···················· ·A·················· ···················· ···················· ···················· ····················
AA 7279 hCoV-19/USA/NY-Wadsworth-237109-01/2020|EPI_ISL_75 ···················· ·A·················· ···················· ···················· ···················· ····················
AA 7280 hCoV-19/USA/NY-Wadsworth-236645-01/2020|EPI_ISL_75 ···················· ···················· ···················· ···················· ···················· ····················
AA 7281 hCoV-19/USA/NY-Wadsworth-236583-01/2020|EPI_ISL_75 ···················· ·············A······ ···················· ···················· ···················· ····················
AA 7282 hCoV-19/USA/NY-Wadsworth-236980-01/2020|EPI_ISL_75 ···················· ·A·················· ···················· ···················· ···················· ····················
AA 7283 hCoV-19/USA/NY-Wadsworth-237073-01/2020|EPI_ISL_75 ···················· ·A·················· ···················· ···················· ···················· ····················
AA 7284 hCoV-19/USA/NY-Wadsworth-237005-01/2020|EPI_ISL_75 ···················· ·A·················· ···················· ···················· ···················· ····················
AA 7285 hCoV-19/USA/NY-Wadsworth-285990-01/2020|EPI_ISL_75 ···················· ···················· ···················· ···················· ···················· ····················
AA 7286 hCoV-19/USA/NY-Wadsworth-270028-01/2020|EPI_ISL_75 ···················· ·A·················· ···················· ···················· ···················· ····················
AA 7287 hCoV-19/USA/NY-Wadsworth-276329-01/2020|EPI_ISL_75 ···················· ···················· ···················· ···················· ···················· ····················
AA 7288 hCoV-19/USA/NY-Wadsworth-277800-01/2020|EPI_ISL_75 ···················· ···················· ···················· ···················· ···················· ····················
AA 7289 hCoV-19/USA/NY-Wadsworth-278122-01/2020|EPI_ISL_75 ···················· ·A·················· ···················· ···················· ···················· ····················
AA 7290 hCoV-19/USA/NY-Wadsworth-278155-01/2020|EPI_ISL_75 ···················· ·A·················· ···················· ···················· ···················· ····················
AA 7291 hCoV-19/USA/NY-Wadsworth-278047-01/2020|EPI_ISL_75 ···················· ·A·················· ···················· ···················· ···················· ····················
AA 7292 hCoV-19/USA/NY-Wadsworth-278048-01/2020|EPI_ISL_75 ···················· ·A·················· ···················· ···················· ···················· ····················
AA 7293 hCoV-19/USA/NY-Wadsworth-278036-01/2020|EPI_ISL_75 ···················· ·A·················· ···················· ···················· ···················· ····················
AA 7294 hCoV-19/USA/NY-Wadsworth-278037-01/2020|EPI_ISL_75 ···················· ·A·················· ···················· ···················· ···················· ····················
AA 7295 hCoV-19/USA/NY-Wadsworth-278038-01/2020|EPI_ISL_75 ···················· ·A·················· ···················· ···················· ···················· ····················

```
AA    7296 hCoV-19/USA/NY-Wadsworth-278039-01/2020|EPI_ISL_75    ····················  ·A··················  ····················  ····················  ····················  ····················
AA    7297 hCoV-19/USA/NY-Wadsworth-278002-01/2020|EPI_ISL_75    ····················  ·A··················  ····················  ····················  ····················  ····················
AA    7298 hCoV-19/Canada/ON-S2016/2020|EPI_ISL_755802|2020-1    ····················  ·A··················  ····················  ····················  ····················  ···A················
AA    7299 hCoV-19/Canada/ON-S2017/2020|EPI_ISL_755803|2020-1    ····················  ·A··················  ····················  ····················  ····················  ···A················
AA    7300 hCoV-19/Canada/ON-S2018/2020|EPI_ISL_755804|2020-1    ····················  ····················  ····················  ····················  ····················  ····················
AA    7301 hCoV-19/Canada/ON-S2019/2020|EPI_ISL_755805|2020-1    ····················  ····················  ····················  ····················  ····················  ····················
AA    7302 hCoV-19/Canada/ON-S2020/2020|EPI_ISL_755806|2020-1    ····················  ····················  ····················  ····················  ····················  ····················
AA    7303 hCoV-19/Canada/ON-S2021/2020|EPI_ISL_755807|2020-1    ····················  ····················  ····················  ····················  ····················  ····················
AA    7304 hCoV-19/Canada/ON-S2022/2020|EPI_ISL_755808|2020-0    ····················  ····················  ····················  ····················  ····················  ····················
AA    7305 hCoV-19/Canada/ON-S2023/2020|EPI_ISL_755809|2020-0    ····················  ····················  ····················  ····················  ····················  ····················
AA    7306 hCoV-19/Canada/ON-S2025/2020|EPI_ISL_755810|2020-1    ····················  ····················  ····················  ····················  ····················  ····················
AA    7307 hCoV-19/Canada/ON-S2027/2020|EPI_ISL_755811|2020-1    ····················  ····················  ····················  ····················  ····················  ····················
AA    7308 hCoV-19/Canada/ON-S2031/2020|EPI_ISL_755812|2020-1    ····················  ·A··················  ····················  ····················  ····················  ···A················
AA    7309 hCoV-19/Canada/ON-S2032/2020|EPI_ISL_755813|2020-1    ····················  ·A··················  ····················  ····················  ····················  ···A················
AA    7310 hCoV-19/Canada/ON-S208/2020|EPI_ISL_755816|2020-03    ····················  ····················  ····················  ····················  ····················  ····················
AA    7311 hCoV-19/Canada/ON-S216/2020|EPI_ISL_755821|2020-03    ····················  ····················  ····················  ····················  ····················  ····················
AA    7312 hCoV-19/Canada/ON-S217/2020|EPI_ISL_755822|2020-03    ····················  ····················  ····················  ····················  ····················  ····················
AA    7313 hCoV-19/Canada/ON-S219/2020|EPI_ISL_755823|2020-03    ····················  ····················  ····················  ····················  ····················  ····················
AA    7314 hCoV-19/Canada/ON-S237/2020|EPI_ISL_755827|2020-03    ····················  ····················  ····················  ····················  ····················  ····················
AA    7315 hCoV-19/Canada/ON-S242/2020|EPI_ISL_755828|2020-03    ····················  ····················  ····················  ····················  ····················  ····················
AA    7316 hCoV-19/Canada/ON-S246/2020|EPI_ISL_755831|2020-03    ····················  ····················  ····················  ····················  ····················  ····················
AA    7317 hCoV-19/Canada/ON-S247/2020|EPI_ISL_755832|2020-03    ····················  ····················  ····················  ····················  ····················  ····················
AA    7318 hCoV-19/Canada/ON-S260/2020|EPI_ISL_755839|2020-03    ····················  ····················  ····················  ····················  ····················  ····················
AA    7319 hCoV-19/Canada/ON-S261/2020|EPI_ISL_755840|2020-03    ····················  ····················  ····················  ····················  ····················  ····················
AA    7320 hCoV-19/Canada/ON-S265/2020|EPI_ISL_755841|2020-03    ····················  ····················  ····················  ····················  ····················  ····················
AA    7321 hCoV-19/Canada/ON-S267/2020|EPI_ISL_755842|2020-03    ····················  ····················  ····················  ····················  ····················  ····················
AA    7322 hCoV-19/Canada/ON-S268/2020|EPI_ISL_755843|2020-03    ····················  ····················  ····················  ····················  ····················  ····················
AA    7323 hCoV-19/Canada/ON-S329/2020|EPI_ISL_755863|2020-03    ····················  ····················  ····················  ····················  ····················  ····················
AA    7324 hCoV-19/Canada/ON-S339/2020|EPI_ISL_755867|2020-03    ····················  ····················  ····················  ····················  ····················  ····················
AA    7325 hCoV-19/Canada/ON-S341/2020|EPI_ISL_755868|2020-03    ····················  ····················  ····················  ····················  ····················  ····················
AA    7326 hCoV-19/Canada/ON-S351/2020|EPI_ISL_755871|2020-03    ····················  ····················  ····················  ····················  ····················  ····················
AA    7327 hCoV-19/Canada/ON-S369/2020|EPI_ISL_755873|2020-03    ····················  ····················  ····················  ····················  ····················  ···A················
AA    7328 hCoV-19/Canada/ON-S370/2020|EPI_ISL_755874|2020-03    ····················  ····················  ····················  ····················  ····················  ····················
AA    7329 hCoV-19/Canada/ON-S381/2020|EPI_ISL_755876|2020-03    ····················  ····················  ····················  ····················  ····················  ····················
AA    7330 hCoV-19/Denmark/DCGC-21617/2020|EPI_ISL_758000|202    ····················  ····················  ····················  ····················  ····················  ····················
AA    7331 hCoV-19/Denmark/DCGC-21638/2020|EPI_ISL_758001|202    ····················  ····················  ····················  ····················  ····················  ····················
AA    7332 hCoV-19/Denmark/DCGC-21645/2020|EPI_ISL_758002|202    ····················  ····················  ····················  ····················  ····················  ····················
AA    7333 hCoV-19/Denmark/DCGC-21655/2020|EPI_ISL_758003|202    ····················  ····················  ····················  ····················  ···C················  ····················
AA    7334 hCoV-19/Denmark/DCGC-21672/2020|EPI_ISL_758004|202    ····················  ····················  ····················  ····················  ····················  ····················
AA    7335 hCoV-19/Denmark/DCGC-21674/2020|EPI_ISL_758005|202    ····················  ····················  ····················  ····················  ····················  ····················
AA    7336 hCoV-19/Denmark/DCGC-21678/2020|EPI_ISL_758006|202    ····················  ····················  ····················  ····················  ····················  ····················
AA    7337 hCoV-19/Denmark/DCGC-21693/2020|EPI_ISL_758007|202    ····················  ····················  ····················  ····················  ····················  ····················
AA    7338 hCoV-19/Denmark/DCGC-21705/2020|EPI_ISL_758008|202    ····················  ····················  ····················  ····················  ····················  ····················
AA    7339 hCoV-19/Denmark/DCGC-21717/2020|EPI_ISL_758009|202    ····················  ····················  ····················  ····················  ····················  ····················
AA    7340 hCoV-19/Denmark/DCGC-21732/2020|EPI_ISL_758010|202    ····················  ····················  ····················  ····················  ····················  ····················
AA    7341 hCoV-19/Denmark/DCGC-21742/2020|EPI_ISL_758011|202    ····················  ····················  ····················  ····················  ····················  ····················
AA    7342 hCoV-19/Denmark/DCGC-21743/2020|EPI_ISL_758012|202    ····················  ····················  ····················  ····················  ···C················  ····················
AA    7343 hCoV-19/Denmark/DCGC-21751/2020|EPI_ISL_758013|202    ····················  ····················  ····················  ····················  ····················  ····················
AA    7344 hCoV-19/Denmark/DCGC-21771/2020|EPI_ISL_758014|202    ····················  ····················  ····················  ····················  ····················  ····················
AA    7345 hCoV-19/Denmark/DCGC-21777/2020|EPI_ISL_758015|202    ····················  ····················  ····················  ····················  ····················  ····················
AA    7346 hCoV-19/Denmark/DCGC-21824/2020|EPI_ISL_758016|202    ····················  ····················  ····················  ····················  ····················  ····················
AA    7347 hCoV-19/Denmark/DCGC-21825/2020|EPI_ISL_758017|202    ····················  ····················  ····················  ····················  ····················  ····················
AA    7348 hCoV-19/Denmark/DCGC-21826/2020|EPI_ISL_758018|202    ····················  ····················  ····················  ····················  ····················  ····················
AA    7349 hCoV-19/Denmark/DCGC-21873/2020|EPI_ISL_758019|202    ····················  ····················  ····················  ····················  ····················  ····················
AA    7350 hCoV-19/Denmark/DCGC-21880/2020|EPI_ISL_758020|202    ····················  ····················  ····················  ····················  ····················  ····················
AA    7351 hCoV-19/Denmark/DCGC-21933/2020|EPI_ISL_758021|202    ····················  ····················  ····················  ····················  ····················  ····················
AA    7352 hCoV-19/Denmark/DCGC-21934/2020|EPI_ISL_758022|202    ····················  ····················  ····················  ····················  ····················  ····················
AA    7353 hCoV-19/Denmark/DCGC-21941/2020|EPI_ISL_758023|202    ····················  ····················  ····················  ····················  ····················  ····················
AA    7354 hCoV-19/Denmark/DCGC-21957/2020|EPI_ISL_758024|202    ····················  ····················  ····················  ····················  ····················  ····················
AA    7355 hCoV-19/Denmark/DCGC-21961/2020|EPI_ISL_758025|202    ····················  ····················  ····················  ····················  ····················  ····················
AA    7356 hCoV-19/Denmark/DCGC-21992/2020|EPI_ISL_758026|202    ····················  ····················  ····················  ····················  ····················  ····················
AA    7357 hCoV-19/Denmark/DCGC-22004/2020|EPI_ISL_758027|202    ····················  ····················  ····················  ····················  ····················  ····················
AA    7358 hCoV-19/Denmark/DCGC-22013/2020|EPI_ISL_758028|202    ····················  ····················  ····················  ····················  ····················  ····················
AA    7359 hCoV-19/Denmark/DCGC-22017/2020|EPI_ISL_758029|202    ····················  ····················  ····················  ····················  ····················  ····················
AA    7360 hCoV-19/Denmark/DCGC-22310/2020|EPI_ISL_758062|202    ····················  ····················  ····················  ····················  ····················  ····················
AA    7361 hCoV-19/Denmark/DCGC-22313/2020|EPI_ISL_758063|202    ····················  ····················  ····················  ····················  ····················  ····················
AA    7362 hCoV-19/Denmark/DCGC-22326/2020|EPI_ISL_758064|202    ····················  ····················  ····················  ····················  ····················  ····················
AA    7363 hCoV-19/Denmark/DCGC-22349/2020|EPI_ISL_758065|202    ····················  ····················  ····················  ····················  ····················  ····················
AA    7364 hCoV-19/Denmark/DCGC-21394/2020|EPI_ISL_758073|202    ····················  ····················  ····················  ····················  ····················  ····················
AA    7365 hCoV-19/Denmark/DCGC-21463/2020|EPI_ISL_758074|202    ····················  ·A··················  ····················  ····················  ····················  ···A················
AA    7366 hCoV-19/Denmark/DCGC-21476/2020|EPI_ISL_758075|202    ····················  ·A··················  ····················  ····················  ····················  ···A················
AA    7367 hCoV-19/Denmark/DCGC-21497/2020|EPI_ISL_758076|202    ····················  ·A··················  ····················  ····················  ····················  ···A················
AA    7368 hCoV-19/Denmark/DCGC-21520/2020|EPI_ISL_758077|202    ····················  ·A··················  ····················  ····················  ····················  ···A················
AA    7369 hCoV-19/Denmark/DCGC-21572/2020|EPI_ISL_758078|202    ····················  ····················  ····················  ····················  ····················  ····················
AA    7370 hCoV-19/Denmark/DCGC-21603/2020|EPI_ISL_758079|202    ····················  ·A··················  ····················  ····················  ····················  ···A················
AA    7371 hCoV-19/Denmark/DCGC-21737/2020|EPI_ISL_758080|202    ····················  ····················  ····················  ····················  ····················  ····················
AA    7372 hCoV-19/Denmark/DCGC-21746/2020|EPI_ISL_758081|202    ····················  ····················  ····················  ····················  ····················  ····················
AA    7373 hCoV-19/Denmark/DCGC-22168/2020|EPI_ISL_758082|202    ····················  ····················  ····················  ····················  ····················  ····················
AA    7374 hCoV-19/Denmark/DCGC-21380/2020|EPI_ISL_758090|202    ····················  ····················  ····················  ····················  ····················  ····················
AA    7375 hCoV-19/Denmark/DCGC-21494/2020|EPI_ISL_758091|202    ····················  ····················  ····················  ····················  ····················  ····················
```

AA 7376 hCoV-19/Denmark/DCGC-21916/2020|EPI_ISL_758092|202 ···················· ···················· ···················· ···················· ···················· ····················
AA 7377 hCoV-19/Denmark/DCGC-22291/2020|EPI_ISL_758093|202 ···················· ···················· ···················· ···················· ···················· ····················
AA 7378 hCoV-19/Denmark/DCGC-20711/2020|EPI_ISL_759320|202 ···················· ···················· ···················· ···················· ···················· ····················
AA 7379 hCoV-19/Denmark/DCGC-20715/2020|EPI_ISL_759321|202 ···················· ···················· ···················· ···················· ···················· ····················
AA 7380 hCoV-19/Denmark/DCGC-20717/2020|EPI_ISL_759322|202 ···················· ···················· ···················· ···················· ···················· ····················
AA 7381 hCoV-19/Denmark/DCGC-20721/2020|EPI_ISL_759323|202 ···················· ···················· ···················· ···················· ···················· ····················
AA 7382 hCoV-19/Denmark/DCGC-20723/2020|EPI_ISL_759324|202 ···················· ···················· ···················· ···················· ···················· ····················
AA 7383 hCoV-19/Denmark/DCGC-20724/2020|EPI_ISL_759325|202 ···················· ···················· ···················· ···················· ···················· ····················
AA 7384 hCoV-19/Denmark/DCGC-20725/2020|EPI_ISL_759326|202 ···················· ···················· ···················· ···················· ···················· ····················
AA 7385 hCoV-19/Denmark/DCGC-20726/2020|EPI_ISL_759327|202 ···················· ···················· ···················· ···················· ···················· ····················
AA 7386 hCoV-19/Denmark/DCGC-20728/2020|EPI_ISL_759328|202 ···················· ···················· ···················· ···················· ···················· ····················
AA 7387 hCoV-19/Denmark/DCGC-20729/2020|EPI_ISL_759329|202 ···················· ···················· ···················· ···················· ···················· ····················
AA 7388 hCoV-19/Denmark/DCGC-20731/2020|EPI_ISL_759330|202 ···················· ···················· ···················· ···················· ···················· ····················
AA 7389 hCoV-19/Denmark/DCGC-20732/2020|EPI_ISL_759331|202 ···················· ···················· ···················· ···················· ···················· ····················
AA 7390 hCoV-19/Denmark/DCGC-20733/2020|EPI_ISL_759332|202 ···················· ···················· ···················· ···················· ···················· ····················
AA 7391 hCoV-19/Denmark/DCGC-20734/2020|EPI_ISL_759333|202 ···················· ···················· ···················· ···················· ···················· ····················
AA 7392 hCoV-19/Denmark/DCGC-20738/2020|EPI_ISL_759334|202 ···················· ·A·················· ···················· ···················· ···················· ···A················
AA 7393 hCoV-19/Denmark/DCGC-20739/2020|EPI_ISL_759335|202 ···················· ···················· ···················· ···················· ···················· ····················
AA 7394 hCoV-19/Denmark/DCGC-20740/2020|EPI_ISL_759336|202 ···················· ···················· ···················· ···················· ···················· ····················
AA 7395 hCoV-19/Denmark/DCGC-20742/2020|EPI_ISL_759337|202 ···················· ·A·················· ···················· ···················· ···················· ···A················
AA 7396 hCoV-19/Denmark/DCGC-20744/2020|EPI_ISL_759338|202 ···················· ···················· ···················· ···················· ···················· ····················
AA 7397 hCoV-19/Denmark/DCGC-20745/2020|EPI_ISL_759339|202 ···················· ···················· ···················· ···················· ···················· ····················
AA 7398 hCoV-19/Denmark/DCGC-20746/2020|EPI_ISL_759340|202 ···················· ···················· ···················· ···················· ···················· ····················
AA 7399 hCoV-19/Denmark/DCGC-20750/2020|EPI_ISL_759341|202 ···················· ···················· ···················· ···················· ···················· ····················
AA 7400 hCoV-19/Denmark/DCGC-20751/2020|EPI_ISL_759342|202 ···················· ···················· ···················· ···················· ···················· ····················
AA 7401 hCoV-19/Denmark/DCGC-20752/2020|EPI_ISL_759343|202 ···················· ···················· ···················· ···················· ···················· ····················
AA 7402 hCoV-19/Denmark/DCGC-20754/2020|EPI_ISL_759344|202 ···················· ···················· ···················· ···················· ···················· ····················
AA 7403 hCoV-19/Denmark/DCGC-20756/2020|EPI_ISL_759345|202 ···················· ···················· ···················· ···················· ···················· ····················
AA 7404 hCoV-19/Denmark/DCGC-20757/2020|EPI_ISL_759346|202 ···················· ···················· ···················· ···················· ···················· ····················
AA 7405 hCoV-19/Denmark/DCGC-20759/2020|EPI_ISL_759347|202 ···················· ···················· ···················· ···················· ···················· ····················
AA 7406 hCoV-19/Denmark/DCGC-20761/2020|EPI_ISL_759348|202 ···················· ···················· ···················· ···················· ···················· ····················
AA 7407 hCoV-19/Denmark/DCGC-20762/2020|EPI_ISL_759349|202 ···················· ···················· ···················· ···················· ···················· ····················
AA 7408 hCoV-19/Denmark/DCGC-20765/2020|EPI_ISL_759350|202 ···················· ···················· ···················· ···················· ···················· ····················
AA 7409 hCoV-19/Denmark/DCGC-20766/2020|EPI_ISL_759351|202 ···················· ···················· ···················· ···················· ···················· ····················
AA 7410 hCoV-19/Denmark/DCGC-20767/2020|EPI_ISL_759352|202 ···················· ···················· ···················· ···················· ···················· ····················
AA 7411 hCoV-19/Denmark/DCGC-20768/2020|EPI_ISL_759353|202 ···················· ···················· ···················· ···················· ···················· ····················
AA 7412 hCoV-19/Denmark/DCGC-20772/2020|EPI_ISL_759354|202 ···················· ···················· ···················· ···················· ···················· ····················
AA 7413 hCoV-19/Denmark/DCGC-20774/2020|EPI_ISL_759355|202 ···················· ···················· ···················· ···················· ···················· ····················
AA 7414 hCoV-19/Denmark/DCGC-20777/2020|EPI_ISL_759356|202 ···················· ···················· ···················· ···················· ···················· ····················
AA 7415 hCoV-19/Denmark/DCGC-20778/2020|EPI_ISL_759357|202 ···················· ···················· ···················· ···················· ···················· ····················
AA 7416 hCoV-19/Denmark/DCGC-20781/2020|EPI_ISL_759358|202 ···················· ···················· ···················· ···················· ···················· ····················
AA 7417 hCoV-19/Denmark/DCGC-20782/2020|EPI_ISL_759359|202 ···················· ···················· ···················· ···················· ···················· ····················
AA 7418 hCoV-19/Denmark/DCGC-20835/2020|EPI_ISL_759393|202 ···················· ···················· ···················· ···················· ···················· ····················
AA 7419 hCoV-19/Denmark/DCGC-20836/2020|EPI_ISL_759394|202 ···················· ···················· ···················· ···················· ···················· ····················
AA 7420 hCoV-19/Denmark/DCGC-20838/2020|EPI_ISL_759395|202 ···················· ···················· ···················· ···················· ···················· ····················
AA 7421 hCoV-19/Denmark/DCGC-20839/2020|EPI_ISL_759396|202 ···················· ····A··············· ···················· ···················· ···················· ····················
AA 7422 hCoV-19/Denmark/DCGC-20840/2020|EPI_ISL_759397|202 ···················· ···················· ···················· ···················· ···················· ····················
AA 7423 hCoV-19/Denmark/DCGC-20841/2020|EPI_ISL_759398|202 ···················· ···················· ···················· ···················· ···················· ····················
AA 7424 hCoV-19/Denmark/DCGC-20842/2020|EPI_ISL_759399|202 ···················· ···················· ···················· ···················· ···················· ····················
AA 7425 hCoV-19/South Korea/KDCA0166/2020|EPI_ISL_747400|2 ···················· ···················· ···················· ···················· ···················· ····················
AA 7426 hCoV-19/South Korea/KDCA0167/2020|EPI_ISL_747401|2 ···················· ···················· ···················· ···················· ···················· ····················
AA 7427 hCoV-19/South Korea/KDCA0168/2020|EPI_ISL_747402|2 ···················· ···················· ···················· ···················· ···················· ····················
AA 7428 hCoV-19/South Korea/KDCA0169/2020|EPI_ISL_747403|2 ···················· ···················· ···················· ···················· ···················· ····················
AA 7429 hCoV-19/South Korea/KDCA0172/2020|EPI_ISL_747406|2 ···················· ···················· ···················· ···················· ···················· ····················
AA 7430 hCoV-19/South Korea/KDCA0173/2020|EPI_ISL_747407|2 ···················· ···················· ···················· ···················· ···················· ····················
AA 7431 hCoV-19/South Korea/KDCA0174/2020|EPI_ISL_747408|2 ···················· ···················· ···················· ···················· ···················· ····················
AA 7432 hCoV-19/South Korea/KDCA0175/2020|EPI_ISL_747409|2 ···················· ···················· ···················· ···················· ···················· ····················
AA 7433 hCoV-19/South Korea/KDCA0176/2020|EPI_ISL_747410|2 ···················· ···················· ···················· ···················· ···················· ····················
AA 7434 hCoV-19/South Korea/KDCA0177/2020|EPI_ISL_747411|2 ···················· ···················· ···················· ···················· ···················· ····················
AA 7435 hCoV-19/South Korea/KDCA0178/2020|EPI_ISL_747412|2 ···················· ···················· ···················· ···················· ···················· ····················
AA 7436 hCoV-19/South Korea/KDCA0179/2020|EPI_ISL_747413|2 ···················· ···················· ···················· ···················· ···················· ····················
AA 7437 hCoV-19/South Korea/KDCA0181/2020|EPI_ISL_747414|2 ···················· ·············A······ ···················· ···················· ···················· ····················
AA 7438 hCoV-19/South Korea/KDCA0183/2020|EPI_ISL_747416|2 ···················· ···················· ···················· ···················· ···················· ····················
AA 7439 hCoV-19/South Korea/KDCA0184/2020|EPI_ISL_747417|2 ···················· ·A·················· ···················· ···················· ···················· ····················
AA 7440 hCoV-19/South Korea/KDCA0185/2020|EPI_ISL_747418|2 ···················· ···················· ···················· ···················· ···················· ····················
AA 7441 hCoV-19/South Korea/KDCA0209/2020|EPI_ISL_747444|2 ···················· ···················· ···················· ···················· ···················· ····················
AA 7442 hCoV-19/South Korea/KDCA0210/2020|EPI_ISL_747445|2 ···················· ···················· ···················· ···················· ···················· ····················
AA 7443 hCoV-19/South Korea/KDCA0211/2020|EPI_ISL_747446|2 ···················· ···················· ···················· ···················· ···················· ····················
AA 7444 hCoV-19/South Korea/KDCA0212/2020|EPI_ISL_747447|2 ···················· ···················· ···················· ···················· ···················· ····················
AA 7445 hCoV-19/South Korea/KDCA0215/2020|EPI_ISL_747450|2 ···················· ···················· ···················· ···················· ···················· ····················
AA 7446 hCoV-19/South Korea/KDCA0216/2020|EPI_ISL_747451|2 ···················· ···················· ···················· ···················· ···················· ····················
AA 7447 hCoV-19/South Korea/KDCA0217/2020|EPI_ISL_747452|2 ···················· ···················· ···················· ···················· ···················· ····················
AA 7448 hCoV-19/South Korea/KDCA0218/2020|EPI_ISL_747453|2 ···················· ···················· ···················· ···················· ···················· ····················
AA 7449 hCoV-19/South Korea/KDCA0219/2020|EPI_ISL_747454|2 ···················· ···················· ···················· ···················· ···················· ····················
AA 7450 hCoV-19/South Korea/KDCA0220/2020|EPI_ISL_747455|2 ···················· ···················· ···················· ···················· ···················· ····················
AA 7451 hCoV-19/South Korea/KDCA0222/2020|EPI_ISL_747456|2 ···················· ···················· ···················· ···················· ···················· ····················
AA 7452 hCoV-19/South Korea/KDCA0225/2020|EPI_ISL_747458|2 ···················· ···················· ···················· ···················· ···················· ····················
AA 7453 hCoV-19/Italy/VEN-IZSVe-20RS2160-29/2020|EPI_ISL_7 ···················· ···················· ···················· ···················· ···················· ····················
AA 7454 hCoV-19/Denmark/DCGC-19017/2020|EPI_ISL_748700|202 ···················· ···················· ···················· ···················· ···················· ····················
AA 7455 hCoV-19/Denmark/DCGC-19018/2020|EPI_ISL_748701|202 ···················· ···················· ···················· ···················· ···················· ····················

| | | | | | | | |
|---|---|---|---|---|---|---|---|
| AA | 7456 hCoV-19/Denmark/DCGC-19019/2020\|EPI_ISL_748702\|202 | ···················· | ···················· | ···················· | ···················· | ···················· | ···················· |
| AA | 7457 hCoV-19/Denmark/DCGC-19020/2020\|EPI_ISL_748703\|202 | ···················· | ·A·················· | ···················· | ···················· | ···················· | ···················· |
| AA | 7458 hCoV-19/Denmark/DCGC-19021/2020\|EPI_ISL_748704\|202 | ···················· | ···················· | ···················· | ···················· | ···················· | ···················· |
| AA | 7459 hCoV-19/Denmark/DCGC-19023/2020\|EPI_ISL_748705\|202 | ···················· | ···················· | ···················· | ···················· | ···················· | ···················· |
| AA | 7460 hCoV-19/Denmark/DCGC-19025/2020\|EPI_ISL_748706\|202 | ···················· | ···················· | ···················· | ···················· | ···················· | ···················· |
| AA | 7461 hCoV-19/Denmark/DCGC-19027/2020\|EPI_ISL_748707\|202 | ···················· | ···················· | ···················· | ···················· | ···················· | ···················· |
| AA | 7462 hCoV-19/Denmark/DCGC-19028/2020\|EPI_ISL_748708\|202 | ···················· | ···················· | ···················· | ···················· | ···················· | ···················· |
| AA | 7463 hCoV-19/Denmark/DCGC-19030/2020\|EPI_ISL_748709\|202 | ···················· | ···················· | ···················· | ···················· | ···················· | ···················· |
| AA | 7464 hCoV-19/Denmark/DCGC-19031/2020\|EPI_ISL_748710\|202 | ···················· | ···················· | ···················· | ···················· | ···················· | ···················· |
| AA | 7465 hCoV-19/Denmark/DCGC-19032/2020\|EPI_ISL_748711\|202 | ···················· | ···················· | ···················· | ···················· | ···················· | ···················· |
| AA | 7466 hCoV-19/Denmark/DCGC-19035/2020\|EPI_ISL_748712\|202 | ···················· | ···················· | ···················· | ···················· | ···················· | ···················· |
| AA | 7467 hCoV-19/Denmark/DCGC-19036/2020\|EPI_ISL_748713\|202 | ···················· | ···················· | ···················· | ···················· | ···················· | ···················· |
| AA | 7468 hCoV-19/Denmark/DCGC-19038/2020\|EPI_ISL_748714\|202 | ···················· | ···················· | ···················· | ···················· | ···················· | ···················· |
| AA | 7469 hCoV-19/Denmark/DCGC-19040/2020\|EPI_ISL_748715\|202 | ···················· | ···················· | ···················· | ···················· | ···················· | ···················· |
| AA | 7470 hCoV-19/Denmark/DCGC-19041/2020\|EPI_ISL_748716\|202 | ···················· | ···················· | ···················· | ···················· | ···················· | ···················· |
| AA | 7471 hCoV-19/Denmark/DCGC-19044/2020\|EPI_ISL_748717\|202 | ···················· | ···················· | ···················· | ···················· | ···················· | ···················· |
| AA | 7472 hCoV-19/Denmark/DCGC-19045/2020\|EPI_ISL_748718\|202 | ···················· | ···················· | ···················· | ···················· | ···················· | ···················· |
| AA | 7473 hCoV-19/Denmark/DCGC-19047/2020\|EPI_ISL_748719\|202 | ···················· | ···················· | ···················· | ···················· | ···················· | ···················· |
| AA | 7474 hCoV-19/Denmark/DCGC-19048/2020\|EPI_ISL_748720\|202 | ···················· | ···················· | ···················· | ···················· | ···················· | ···················· |
| AA | 7475 hCoV-19/Denmark/DCGC-19050/2020\|EPI_ISL_748721\|202 | ···················· | ···················· | ···················· | ···················· | ···················· | ···················· |
| AA | 7476 hCoV-19/Denmark/DCGC-19051/2020\|EPI_ISL_748722\|202 | ···················· | ···················· | ···················· | ···················· | ···················· | ···················· |
| AA | 7477 hCoV-19/Denmark/DCGC-19052/2020\|EPI_ISL_748723\|202 | ···················· | ···················· | ···················· | ···················· | ···················· | ···················· |
| AA | 7478 hCoV-19/Denmark/DCGC-19053/2020\|EPI_ISL_748724\|202 | ···················· | ···················· | ···················· | ···················· | ···················· | ···················· |
| AA | 7479 hCoV-19/Denmark/DCGC-19054/2020\|EPI_ISL_748725\|202 | ···················· | ···················· | ···················· | ···················· | ···················· | ···················· |
| AA | 7480 hCoV-19/Denmark/DCGC-19055/2020\|EPI_ISL_748726\|202 | ···················· | ···················· | ···················· | ···················· | ···················· | ···················· |
| AA | 7481 hCoV-19/Denmark/DCGC-19059/2020\|EPI_ISL_748727\|202 | ···················· | ···················· | ···················· | ···················· | ···················· | ···················· |
| AA | 7482 hCoV-19/Denmark/DCGC-19060/2020\|EPI_ISL_748728\|202 | ···················· | ···················· | ···················· | ···················· | ···················· | ···················· |
| AA | 7483 hCoV-19/Denmark/DCGC-19061/2020\|EPI_ISL_748729\|202 | ···················· | ···················· | ···················· | ···················· | ···················· | ···················· |
| AA | 7484 hCoV-19/Denmark/DCGC-19064/2020\|EPI_ISL_748730\|202 | ···················· | ···················· | ···················· | ···················· | ···················· | ···················· |
| AA | 7485 hCoV-19/Denmark/DCGC-19065/2020\|EPI_ISL_748731\|202 | ···················· | ···················· | ···················· | ···················· | ···················· | ···················· |
| AA | 7486 hCoV-19/Denmark/DCGC-19066/2020\|EPI_ISL_748732\|202 | ···················· | ···················· | ···················· | ···················· | ···················· | ···················· |
| AA | 7487 hCoV-19/Denmark/DCGC-19067/2020\|EPI_ISL_748733\|202 | ···················· | ···················· | ···················· | ···················· | ···················· | ···················· |
| AA | 7488 hCoV-19/Denmark/DCGC-19069/2020\|EPI_ISL_748734\|202 | ···················· | ···················· | ···················· | ···················· | ···················· | ···················· |
| AA | 7489 hCoV-19/Denmark/DCGC-19070/2020\|EPI_ISL_748735\|202 | ···················· | ···················· | ···················· | ···················· | ···················· | ···················· |
| AA | 7490 hCoV-19/Denmark/DCGC-19072/2020\|EPI_ISL_748736\|202 | ···················· | ···················· | ···················· | ···················· | ···················· | ···················· |
| AA | 7491 hCoV-19/Denmark/DCGC-19073/2020\|EPI_ISL_748737\|202 | ···················· | ···················· | ···················· | ···················· | ···················· | ···················· |
| AA | 7492 hCoV-19/Denmark/DCGC-19074/2020\|EPI_ISL_748738\|202 | ···················· | ···················· | ···················· | ···················· | ···················· | ···················· |
| AA | 7493 hCoV-19/Denmark/DCGC-19075/2020\|EPI_ISL_748739\|202 | ···················· | ···················· | ···················· | ···················· | ···················· | ···················· |
| AA | 7494 hCoV-19/Denmark/DCGC-19076/2020\|EPI_ISL_748740\|202 | ···················· | ···················· | ···················· | ···················· | ···················· | ···················· |
| AA | 7495 hCoV-19/Denmark/DCGC-19078/2020\|EPI_ISL_748741\|202 | ···················· | ·A·················· | ···················· | ···················· | ···················· | ···A················ |
| AA | 7496 hCoV-19/Denmark/DCGC-19079/2020\|EPI_ISL_748742\|202 | ···················· | ···················· | ···················· | ···················· | ···················· | ···················· |
| AA | 7497 hCoV-19/Denmark/DCGC-19080/2020\|EPI_ISL_748743\|202 | ···················· | ···················· | ···················· | ···················· | ···················· | ···················· |
| AA | 7498 hCoV-19/Denmark/DCGC-19081/2020\|EPI_ISL_748744\|202 | ···················· | ···················· | ···················· | ···················· | ···················· | ···················· |
| AA | 7499 hCoV-19/Denmark/DCGC-19082/2020\|EPI_ISL_748745\|202 | ···················· | ···················· | ···················· | ···················· | ···················· | ···················· |
| AA | 7500 hCoV-19/Denmark/DCGC-19084/2020\|EPI_ISL_748746\|202 | ···················· | ···················· | ···················· | ···················· | ······T············· | ···················· |
| AA | 7501 hCoV-19/Denmark/DCGC-19085/2020\|EPI_ISL_748747\|202 | ···················· | ···················· | ···················· | ···················· | ···················· | ···················· |
| AA | 7502 hCoV-19/Denmark/DCGC-19086/2020\|EPI_ISL_748748\|202 | ···················· | ···················· | ···················· | ···················· | ···················· | ···················· |
| AA | 7503 hCoV-19/Denmark/DCGC-19087/2020\|EPI_ISL_748749\|202 | ···················· | ···················· | ···················· | ···················· | ···················· | ···················· |
| AA | 7504 hCoV-19/Denmark/DCGC-19144/2020\|EPI_ISL_748779\|202 | ···················· | ···················· | ···················· | ···················· | ···················· | ···················· |
| AA | 7505 hCoV-19/Denmark/DCGC-19147/2020\|EPI_ISL_748780\|202 | ···················· | ···················· | ···················· | ···················· | ···················· | ···················· |
| AA | 7506 hCoV-19/Denmark/DCGC-19148/2020\|EPI_ISL_748781\|202 | ···················· | ···················· | ···················· | ···················· | ···················· | ···················· |
| AA | 7507 hCoV-19/Denmark/DCGC-19152/2020\|EPI_ISL_748782\|202 | ···················· | ···················· | ···················· | ···················· | ···················· | ···················· |
| AA | 7508 hCoV-19/Denmark/DCGC-19153/2020\|EPI_ISL_748783\|202 | ···················· | ···················· | ···················· | ···················· | ···················· | ···················· |
| AA | 7509 hCoV-19/Denmark/DCGC-19155/2020\|EPI_ISL_748784\|202 | ···················· | ···················· | ···················· | ···················· | ···················· | ···················· |
| AA | 7510 hCoV-19/Denmark/DCGC-19156/2020\|EPI_ISL_748785\|202 | ···················· | ···················· | ···················· | ···················· | ···················· | ···················· |
| AA | 7511 hCoV-19/Denmark/DCGC-19157/2020\|EPI_ISL_748786\|202 | ···················· | ·A·················· | ···················· | ···················· | ···················· | ···················· |
| AA | 7512 hCoV-19/Denmark/DCGC-19162/2020\|EPI_ISL_748787\|202 | ···················· | ···················· | ···················· | ···················· | ···················· | ···················· |
| AA | 7513 hCoV-19/Denmark/DCGC-19163/2020\|EPI_ISL_748788\|202 | ···················· | ···················· | ···················· | ···················· | ···················· | ···················· |
| AA | 7514 hCoV-19/Denmark/DCGC-19166/2020\|EPI_ISL_748789\|202 | ···················· | ···················· | ···················· | ···················· | ···················· | ···················· |
| AA | 7515 hCoV-19/USA/HI-H200694/2020\|EPI_ISL_753216\|2020-12 | ···················· | ·A·················· | ···················· | ···················· | ···················· | ···················· |
| AA | 7516 hCoV-19/USA/HI-H200695/2020\|EPI_ISL_753217\|2020-12 | ···················· | ·A·················· | ···················· | ···················· | ···················· | ···················· |
| AA | 7517 hCoV-19/USA/HI-H200696/2020\|EPI_ISL_753218\|2020-12 | ···················· | ·A·················· | ···················· | ···················· | ···················· | ···················· |
| AA | 7518 hCoV-19/USA/HI-H200698/2020\|EPI_ISL_753219\|2020-12 | ···················· | ·A·················· | ···················· | ···················· | ···················· | ···················· |
| AA | 7519 hCoV-19/USA/HI-H200699/2020\|EPI_ISL_753220\|2020-12 | ···················· | ·A·················· | ···················· | ···················· | ···················· | ···················· |
| AA | 7520 hCoV-19/USA/HI-H200700/2020\|EPI_ISL_753221\|2020-11 | ···················· | ·A·················· | ···················· | ···················· | ···················· | ···················· |
| AA | 7521 hCoV-19/USA/HI-H200701/2020\|EPI_ISL_753222\|2020-11 | ···················· | ·A·················· | ···················· | ···················· | ···················· | ···················· |
| AA | 7522 hCoV-19/USA/HI-H200702/2020\|EPI_ISL_753223\|2020-12 | ···················· | ·A·················· | ···················· | ···················· | ···················· | ···················· |
| AA | 7523 hCoV-19/USA/HI-H200703/2020\|EPI_ISL_753224\|2020-12 | ···················· | ·A·················· | ···················· | ···················· | ···················· | ···················· |
| AA | 7524 hCoV-19/USA/HI-H200704/2020\|EPI_ISL_753225\|2020-12 | ···················· | ···················· | ···················· | ···················· | ···················· | ···················· |
| AA | 7525 hCoV-19/USA/HI-H200705/2020\|EPI_ISL_753226\|2020-12 | ···················· | ·A·················· | ···················· | ···················· | ···················· | ···················· |
| AA | 7526 hCoV-19/USA/HI-H200706/2020\|EPI_ISL_753227\|2020-12 | ···················· | ·A·················· | ···················· | ···················· | ···················· | ···················· |
| AA | 7527 hCoV-19/USA/HI-H200707/2020\|EPI_ISL_753228\|2020-12 | ···················· | ·A·················· | ···················· | ···················· | ···················· | ···················· |
| AA | 7528 hCoV-19/USA/HI-H200708/2020\|EPI_ISL_753229\|2020-12 | ···················· | ·A·················· | ···················· | ···················· | ···················· | ···················· |
| AA | 7529 hCoV-19/USA/HI-H200709/2020\|EPI_ISL_753230\|2020-12 | ···················· | ·A·················· | ···················· | ···················· | ···················· | ···················· |
| AA | 7530 hCoV-19/USA/HI-H200710/2020\|EPI_ISL_753231\|2020-12 | ···················· | ·A·················· | ···················· | ···················· | ···················· | ···················· |
| AA | 7531 hCoV-19/USA/HI-H200711/2020\|EPI_ISL_753232\|2020-12 | ···················· | ·A·················· | ···················· | ···················· | ···················· | ···················· |
| AA | 7532 hCoV-19/USA/HI-H200712/2020\|EPI_ISL_753233\|2020-12 | ···················· | ·A·················· | ···················· | ···················· | ···················· | ···················· |
| AA | 7533 hCoV-19/USA/HI-H200713/2020\|EPI_ISL_753234\|2020-12 | ···················· | ·A·················· | ···················· | ···················· | ···················· | ···················· |
| AA | 7534 hCoV-19/USA/HI-H200714/2020\|EPI_ISL_753235\|2020-12 | ···················· | ·A·················· | ···················· | ···················· | ···················· | ···················· |
| AA | 7535 hCoV-19/USA/HI-H200715/2020\|EPI_ISL_753236\|2020-12 | ···················· | ·A·················· | ···················· | ···················· | ···················· | ···················· |

| | | | | | | | | |
|---|---|---|---|---|---|---|---|---|
| AA | 7536 | hCoV-19/USA/HI-H200716/2020\|EPI_ISL_753237\|2020-12 | ···················· | ·A·················· | ···················· | ···················· | ···················· | ···················· |
| AA | 7537 | hCoV-19/USA/HI-H200717/2020\|EPI_ISL_753238\|2020-12 | ···················· | ·A·················· | ···················· | ···················· | ···················· | ···················· |
| AA | 7538 | hCoV-19/USA/HI-H200718/2020\|EPI_ISL_753239\|2020-12 | ···················· | ·A·················· | ···················· | ···················· | ···················· | ···················· |
| AA | 7539 | hCoV-19/USA/HI-H200719/2020\|EPI_ISL_753240\|2020-12 | ···················· | ·A·················· | ···················· | ···················· | ···················· | ···················· |
| AA | 7540 | hCoV-19/USA/HI-H200720/2020\|EPI_ISL_753241\|2020-12 | ···················· | ···················· | ···················· | ···················· | ···················· | ···················· |
| AA | 7541 | hCoV-19/USA/HI-H200721/2020\|EPI_ISL_753242\|2020-12 | ···················· | ·A·················· | ···················· | ···················· | ···················· | ···················· |
| AA | 7542 | hCoV-19/USA/HI-H200722/2020\|EPI_ISL_753243\|2020-12 | ···················· | ·A·················· | ···················· | ···················· | ···················· | ···················· |
| AA | 7543 | hCoV-19/USA/HI-H200723/2020\|EPI_ISL_753244\|2020-12 | ···················· | ·A·················· | ···················· | ···················· | ···················· | ···················· |
| AA | 7544 | hCoV-19/USA/HI-H200724/2020\|EPI_ISL_753245\|2020-12 | ···················· | ·A·················· | ···················· | ···················· | ···················· | ···················· |
| AA | 7545 | hCoV-19/USA/HI-H200725/2020\|EPI_ISL_753246\|2020-12 | ···················· | ·A·················· | ···················· | ···················· | ···················· | ···················· |
| AA | 7546 | hCoV-19/USA/HI-H200726/2020\|EPI_ISL_753247\|2020-12 | ···················· | ·A·················· | ···················· | ···················· | ···················· | ···················· |
| AA | 7547 | hCoV-19/USA/HI-H200727/2020\|EPI_ISL_753248\|2020-12 | ···················· | ·A·················· | ···················· | ···················· | ···················· | ···················· |
| AA | 7548 | hCoV-19/USA/HI-H200728/2020\|EPI_ISL_753249\|2020-12 | ···················· | ·A·················· | ···················· | ···················· | ···················· | ···················· |
| AA | 7549 | hCoV-19/USA/HI-H200729/2020\|EPI_ISL_753250\|2020-12 | ···················· | ·A·················· | ···················· | ···················· | ···················· | ···················· |
| AA | 7550 | hCoV-19/USA/HI-H200730/2020\|EPI_ISL_753251\|2020-12 | ···················· | ·A·················· | ···················· | ···················· | ···················· | ···················· |
| AA | 7551 | hCoV-19/USA/HI-H200731/2020\|EPI_ISL_753252\|2020-12 | ···················· | ·A·················· | ···················· | ···················· | ···················· | ···················· |
| AA | 7552 | hCoV-19/USA/HI-H200732/2020\|EPI_ISL_753253\|2020-12 | ···················· | ·A·················· | ···················· | ···················· | ···················· | ···················· |
| AA | 7553 | hCoV-19/USA/HI-H200733/2020\|EPI_ISL_753254\|2020-12 | ···················· | ·A·················· | ···················· | ···················· | ···················· | ···················· |
| AA | 7554 | hCoV-19/USA/HI-H200734/2020\|EPI_ISL_753255\|2020-12 | ···················· | ·A·················· | ···················· | ···················· | ···················· | ···················· |
| AA | 7555 | hCoV-19/USA/HI-H200735/2020\|EPI_ISL_753256\|2020-12 | ···················· | ·A·················· | ···················· | ···················· | ···················· | ···················· |
| AA | 7556 | hCoV-19/USA/HI-H200736/2020\|EPI_ISL_753257\|2020-12 | ···················· | ·A·················· | ···················· | ···················· | ···················· | ···················· |
| AA | 7557 | hCoV-19/USA/HI-H200737/2020\|EPI_ISL_753258\|2020-12 | ···················· | ·A·················· | ···················· | ···················· | ···················· | ···················· |
| AA | 7558 | hCoV-19/USA/HI-H200738/2020\|EPI_ISL_753259\|2020-12 | ···················· | ·A·················· | ···················· | ···················· | ···················· | ···················· |
| AA | 7559 | hCoV-19/USA/CA-CHLA-PLM75672420/2020\|EPI_ISL_75327 | ···················· | ···················· | ···················· | ···················· | ···················· | ···················· |
| AA | 7560 | hCoV-19/USA/CA-CHLA-PLM24203810/2020\|EPI_ISL_75327 | ···················· | ·A·················· | ···················· | ···················· | ···················· | ···················· |
| AA | 7561 | hCoV-19/USA/CA-CHLA-PLM87208720/2020\|EPI_ISL_75327 | ···················· | ···················· | ···················· | ···················· | ···················· | ···················· |
| AA | 7562 | hCoV-19/USA/CA-CHLA-PLM42921945/2020\|EPI_ISL_75327 | ···················· | ·A·················· | ···················· | ···················· | ···················· | ···················· |
| AA | 7563 | hCoV-19/USA/CA-CHLA-PLM69614617/2020\|EPI_ISL_75327 | ···················· | ···················· | ···················· | ···················· | ···················· | ···················· |
| AA | 7564 | hCoV-19/USA/CA-CHLA-PLM42759950/2020\|EPI_ISL_75327 | ···················· | ·A·················· | ···················· | ···················· | ···················· | ···················· |
| AA | 7565 | hCoV-19/USA/CA-CHLA-PLM02264454/2020\|EPI_ISL_75328 | ···················· | ···················· | ···················· | ···················· | ···················· | ···················· |
| AA | 7566 | hCoV-19/USA/CA-CHLA-PLM24227074/2020\|EPI_ISL_75328 | ···················· | ···················· | ···················· | ···················· | ···················· | ···················· |
| AA | 7567 | hCoV-19/USA/CA-CHLA-PLM31973561/2020\|EPI_ISL_75328 | ···················· | ···················· | ···················· | ···················· | ···················· | ···················· |
| AA | 7568 | hCoV-19/USA/CA-CHLA-PLM70050531/2020\|EPI_ISL_75328 | ···················· | ···················· | ···················· | ···················· | ···················· | ···················· |
| AA | 7569 | hCoV-19/USA/CA-CHLA-PLM24319824/2020\|EPI_ISL_75328 | ···················· | ·A·················· | ···················· | ···················· | ···················· | ···················· |
| AA | 7570 | hCoV-19/USA/CA-CHLA-PLM01113476/2020\|EPI_ISL_75328 | ···················· | ···················· | ···················· | ···················· | ···················· | ···················· |
| AA | 7571 | hCoV-19/USA/CA-CHLA-PLM73121337/2020\|EPI_ISL_75328 | ···················· | ···················· | ···················· | ···················· | ···················· | ···················· |
| AA | 7572 | hCoV-19/USA/CA-CHLA-PLM45535740/2020\|EPI_ISL_75328 | ···················· | ···················· | ···················· | ···················· | ···················· | ···················· |
| AA | 7573 | hCoV-19/USA/CA-CHLA-PLM55663009/2020\|EPI_ISL_75328 | ···················· | ···················· | ···················· | ···················· | ···················· | ···················· |
| AA | 7574 | hCoV-19/USA/CA-CHLA-PLM83454072/2020\|EPI_ISL_75328 | ···················· | ·A·················· | ···················· | ···················· | ···················· | ···················· |
| AA | 7575 | hCoV-19/USA/CA-CHLA-PLM61223149/2020\|EPI_ISL_75329 | ···················· | ···················· | ···················· | ···················· | ···················· | ···················· |
| AA | 7576 | hCoV-19/USA/CA-CHLA-PLM47560065/2020\|EPI_ISL_75329 | ···················· | ···················· | ···················· | ···················· | ···················· | ···················· |
| AA | 7577 | hCoV-19/USA/CA-CHLA-PLM29698897/2020\|EPI_ISL_75329 | ···················· | ···················· | ···················· | ···················· | ···················· | ···················· |
| AA | 7578 | hCoV-19/USA/CA-CHLA-PLM94026245/2020\|EPI_ISL_75329 | ···················· | ···················· | ···················· | ···················· | ···················· | ···················· |
| AA | 7579 | hCoV-19/USA/CA-CHLA-PLM28873007/2020\|EPI_ISL_75329 | ···················· | ···················· | ···················· | ···················· | ···················· | ···················· |
| AA | 7580 | hCoV-19/USA/CA-CHLA-PLM82577071/2020\|EPI_ISL_75329 | ···················· | ···················· | ···················· | ···················· | ···················· | ···················· |
| AA | 7581 | hCoV-19/USA/CA-CHLA-PLM32753781/2020\|EPI_ISL_75329 | ···················· | ···················· | ···················· | ···················· | ···················· | ···················· |
| AA | 7582 | hCoV-19/USA/CA-CHLA-PLM32957810/2020\|EPI_ISL_75329 | ···················· | ···················· | ···················· | ···················· | ···················· | ···················· |
| AA | 7583 | hCoV-19/USA/CA-CHLA-PLM81394288/2020\|EPI_ISL_75329 | ···················· | ···················· | ···················· | ···················· | ···················· | ···················· |
| AA | 7584 | hCoV-19/USA/CA-CHLA-PLM65167224/2020\|EPI_ISL_75329 | ···················· | ···················· | ···················· | ···················· | ···················· | ···················· |
| AA | 7585 | hCoV-19/USA/NY-Wadsworth-285711-01/2020\|EPI_ISL_75 | ···················· | ···················· | ···················· | ···················· | ···················· | ···················· |
| AA | 7586 | hCoV-19/USA/NY-Wadsworth-285721-01/2020\|EPI_ISL_75 | ···················· | ···················· | ···················· | ···················· | ···················· | ···················· |
| AA | 7587 | hCoV-19/USA/NY-Wadsworth-285732-01/2020\|EPI_ISL_75 | ···················· | ···················· | ···················· | ···················· | ···················· | ···················· |
| AA | 7588 | hCoV-19/USA/NY-Wadsworth-286011-01/2020\|EPI_ISL_75 | ···················· | ···················· | ···················· | ···················· | ···················· | ···················· |
| AA | 7589 | hCoV-19/USA/NY-Wadsworth-286012-01/2020\|EPI_ISL_75 | ···················· | ···················· | ···················· | ···················· | ···················· | ···················· |
| AA | 7590 | hCoV-19/USA/NY-Wadsworth-286014-01/2020\|EPI_ISL_75 | ···················· | ···················· | ···················· | ···················· | ···················· | ···················· |
| AA | 7591 | hCoV-19/USA/NY-Wadsworth-286020-01/2020\|EPI_ISL_75 | ···················· | ···················· | ···················· | ···················· | ···················· | ···················· |
| AA | 7592 | hCoV-19/USA/NY-Wadsworth-286021-01/2020\|EPI_ISL_75 | ···················· | ···················· | ···················· | ···················· | ···················· | ···················· |
| AA | 7593 | hCoV-19/USA/NY-Wadsworth-285896-01/2020\|EPI_ISL_75 | ···················· | ···················· | ···················· | ···················· | ···················· | ···················· |
| AA | 7594 | hCoV-19/USA/NY-Wadsworth-286048-01/2020\|EPI_ISL_75 | ···················· | ···················· | ···················· | ···················· | ···················· | ···················· |
| AA | 7595 | hCoV-19/USA/NY-Wadsworth-286049-01/2020\|EPI_ISL_75 | ···················· | ···················· | ···················· | ···················· | ···················· | ···················· |
| AA | 7596 | hCoV-19/USA/NY-Wadsworth-286057-01/2020\|EPI_ISL_75 | ···················· | ···················· | ···················· | ···················· | ···················· | ···················· |
| AA | 7597 | hCoV-19/USA/NY-Wadsworth-286058-01/2020\|EPI_ISL_75 | ···················· | ···················· | ···················· | ···················· | ···················· | ···················· |
| AA | 7598 | hCoV-19/USA/NY-Wadsworth-286294-01/2020\|EPI_ISL_75 | ···················· | ···················· | ···················· | ···················· | ···················· | ···················· |
| AA | 7599 | hCoV-19/USA/NY-Wadsworth-286233-01/2020\|EPI_ISL_75 | ···················· | ·A·················· | ···················· | ···················· | ···················· | ···················· |
| AA | 7600 | hCoV-19/USA/NY-Wadsworth-286242-01/2020\|EPI_ISL_75 | ···················· | ·A·················· | ···················· | ···················· | ···················· | ···················· |
| AA | 7601 | hCoV-19/USA/NY-Wadsworth-286243-01/2020\|EPI_ISL_75 | ···················· | ·A·················· | ···················· | ···················· | ···················· | ···················· |
| AA | 7602 | hCoV-19/USA/NY-Wadsworth-286244-01/2020\|EPI_ISL_75 | ···················· | ·A·················· | ···················· | ···················· | ···················· | ···················· |
| AA | 7603 | hCoV-19/USA/NY-Wadsworth-286247-01/2020\|EPI_ISL_75 | ···················· | ·A·················· | ···················· | ···················· | ···················· | ···················· |
| AA | 7604 | hCoV-19/USA/NY-Wadsworth-286249-01/2020\|EPI_ISL_75 | ···················· | ·A·················· | ···················· | ···················· | ···················· | ···················· |
| AA | 7605 | hCoV-19/USA/NY-Wadsworth-286251-01/2020\|EPI_ISL_75 | ···················· | ···················· | ···················· | ···················· | ···················· | ···················· |
| AA | 7606 | hCoV-19/USA/NY-Wadsworth-286122-01/2020\|EPI_ISL_75 | ···················· | ·A·················· | ···················· | ···················· | ···················· | ···················· |
| AA | 7607 | hCoV-19/USA/NY-Wadsworth-286307-01/2020\|EPI_ISL_75 | ···················· | ···················· | ···················· | ···················· | ···················· | ···················· |
| AA | 7608 | hCoV-19/USA/NY-Wadsworth-286088-01/2020\|EPI_ISL_75 | ···················· | ···················· | ···················· | ···················· | T··················· | ···················· |
| AA | 7609 | hCoV-19/USA/NY-Wadsworth-286312-01/2020\|EPI_ISL_75 | ···················· | ···················· | ···················· | ···················· | ···················· | ···················· |
| AA | 7610 | hCoV-19/USA/NY-Wadsworth-285736-01/2020\|EPI_ISL_75 | ···················· | ·A·················· | ···················· | ···················· | ···················· | ···················· |
| AA | 7611 | hCoV-19/USA/NY-Wadsworth-286380-01/2020\|EPI_ISL_75 | ···················· | ···················· | ···················· | ···················· | ···················· | ···················· |
| AA | 7612 | hCoV-19/USA/NY-Wadsworth-286381-01/2020\|EPI_ISL_75 | ···················· | ···················· | ···················· | ···················· | ···················· | ···················· |
| AA | 7613 | hCoV-19/USA/NY-Wadsworth-269500-01/2020\|EPI_ISL_75 | ···················· | ···················· | ···················· | ···················· | ···················· | ···················· |
| AA | 7614 | hCoV-19/USA/NY-Wadsworth-269507-01/2020\|EPI_ISL_75 | ···················· | ···················· | ···················· | ···················· | ···················· | ···················· |
| AA | 7615 | hCoV-19/USA/NY-Wadsworth-275044-01/2020\|EPI_ISL_75 | ···················· | ···················· | ···················· | ···················· | ···················· | ···················· |

| | | | | | | | | |
|---|---|---|---|---|---|---|---|---|
| AA | 7616 | hCoV-19/USA/NY-Wadsworth-274786-01/2020\|EPI_ISL_75 | ···················· | ···················· | ···················· | ···················· | ···················· | ···················· |
| AA | 7617 | hCoV-19/USA/NY-Wadsworth-275442-01/2020\|EPI_ISL_75 | ···················· | ·A·················· | ···················· | ···················· | ···················· | ···················· |
| AA | 7618 | hCoV-19/USA/NY-Wadsworth-275445-01/2020\|EPI_ISL_75 | ···················· | ·A·················· | ···················· | ···················· | ···················· | ···················· |
| AA | 7619 | hCoV-19/USA/NY-Wadsworth-276936-01/2020\|EPI_ISL_75 | ···················· | ·A·················· | ···················· | ···················· | ···················· | ···················· |
| AA | 7620 | hCoV-19/USA/NY-Wadsworth-276906-01/2020\|EPI_ISL_75 | ···················· | ···················· | ···················· | ···················· | ···················· | ···················· |
| AA | 7621 | hCoV-19/USA/NY-Wadsworth-276659-01/2020\|EPI_ISL_75 | ···················· | ···················· | ···················· | ···················· | ···················· | ···················· |
| AA | 7622 | hCoV-19/USA/NY-Wadsworth-277233-01/2020\|EPI_ISL_75 | ···················· | ···················· | ···················· | ···················· | ···················· | ···················· |
| AA | 7623 | hCoV-19/USA/NY-Wadsworth-277184-01/2020\|EPI_ISL_75 | ···················· | ···················· | ···················· | ···················· | ···················· | ···················· |
| AA | 7624 | hCoV-19/USA/NY-Wadsworth-277125-01/2020\|EPI_ISL_75 | ···················· | ···················· | ···················· | ···················· | ···················· | ···················· |
| AA | 7625 | hCoV-19/USA/NY-Wadsworth-277726-01/2020\|EPI_ISL_75 | ···················· | ···················· | ···················· | ···················· | ···················· | ···················· |
| AA | 7626 | hCoV-19/USA/NY-Wadsworth-277729-01/2020\|EPI_ISL_75 | ···················· | ···················· | ···················· | ···················· | ···················· | ···················· |
| AA | 7627 | hCoV-19/USA/NY-Wadsworth-277731-01/2020\|EPI_ISL_75 | ···················· | ···················· | ···················· | ···················· | ···················· | ···················· |
| AA | 7628 | hCoV-19/Canada/ON-S397/2020\|EPI_ISL_755880\|2020-03 | ···················· | ···················· | ···················· | ···················· | ···················· | ···················· |
| AA | 7629 | hCoV-19/Canada/ON-S1494/2020\|EPI_ISL_755883\|2020-0 | ···················· | ···················· | ···················· | ···················· | ···················· | ···················· |
| AA | 7630 | hCoV-19/Canada/ON-S1495/2020\|EPI_ISL_755884\|2020-0 | ···················· | ···················· | ···················· | ···················· | ···················· | ···················· |
| AA | 7631 | hCoV-19/Canada/ON-S1496/2020\|EPI_ISL_755885\|2020-0 | ···················· | ···················· | ···················· | ···················· | ···················· | ···················· |
| AA | 7632 | hCoV-19/Canada/ON-S1499/2020\|EPI_ISL_755888\|2020-0 | ···················· | ·A·················· | ···················· | ···················· | ···················· | ··A················· |
| AA | 7633 | hCoV-19/Canada/ON-S1501/2020\|EPI_ISL_755890\|2020-0 | ···················· | ···················· | ···················· | ···················· | ···················· | ···················· |
| AA | 7634 | hCoV-19/Canada/ON-S1504/2020\|EPI_ISL_755892\|2020-0 | ···················· | ···················· | ···················· | ···················· | ···················· | ···················· |
| AA | 7635 | hCoV-19/Canada/ON-S1512/2020\|EPI_ISL_755895\|2020-0 | ···················· | ···················· | ···················· | ···················· | ···················· | ···················· |
| AA | 7636 | hCoV-19/Canada/ON-S1513/2020\|EPI_ISL_755896\|2020-0 | ···················· | ·A·················· | ···················· | ···················· | ···················· | ···················· |
| AA | 7637 | hCoV-19/Canada/ON-S1514/2020\|EPI_ISL_755897\|2020-0 | ···················· | ···················· | ···················· | ···················· | ···················· | ···················· |
| AA | 7638 | hCoV-19/Canada/ON-S1517/2020\|EPI_ISL_755899\|2020-0 | ···················· | ···················· | ···················· | ···················· | ···················· | ···················· |
| AA | 7639 | hCoV-19/Denmark/DCGC-21565/2020\|EPI_ISL_758103\|202 | ···················· | ···················· | ···················· | ···················· | ···················· | ···················· |
| AA | 7640 | hCoV-19/Denmark/DCGC-21590/2020\|EPI_ISL_758104\|202 | ···················· | ···················· | ···················· | ···················· | ···················· | ···················· |
| AA | 7641 | hCoV-19/Denmark/DCGC-20843/2020\|EPI_ISL_759400\|202 | ···················· | ···················· | ···················· | ···················· | ···················· | ···················· |
| AA | 7642 | hCoV-19/Denmark/DCGC-20844/2020\|EPI_ISL_759401\|202 | ···················· | ···················· | ···················· | ···················· | ···················· | ···················· |
| AA | 7643 | hCoV-19/Denmark/DCGC-20846/2020\|EPI_ISL_759402\|202 | ···················· | ···················· | ···················· | ···················· | ···················· | ···················· |
| AA | 7644 | hCoV-19/Denmark/DCGC-20847/2020\|EPI_ISL_759403\|202 | ···················· | ···················· | ···················· | ···················· | ···················· | ···················· |
| AA | 7645 | hCoV-19/Denmark/DCGC-20848/2020\|EPI_ISL_759404\|202 | ···················· | ···················· | ···················· | ···················· | ···················· | ···················· |
| AA | 7646 | hCoV-19/Denmark/DCGC-20849/2020\|EPI_ISL_759405\|202 | ···················· | ···················· | ···················· | ···················· | ···················· | ···················· |
| AA | 7647 | hCoV-19/Denmark/DCGC-20850/2020\|EPI_ISL_759406\|202 | ···················· | ···················· | ···················· | ···················· | ···················· | ···················· |
| AA | 7648 | hCoV-19/Denmark/DCGC-20851/2020\|EPI_ISL_759407\|202 | ···················· | ···················· | ···················· | ···················· | ···················· | ···················· |
| AA | 7649 | hCoV-19/Denmark/DCGC-20852/2020\|EPI_ISL_759408\|202 | ···················· | ···················· | ···················· | ···················· | ···················· | ···················· |
| AA | 7650 | hCoV-19/Denmark/DCGC-20853/2020\|EPI_ISL_759409\|202 | ···················· | ···················· | ···················· | ···················· | ···················· | ···················· |
| AA | 7651 | hCoV-19/Denmark/DCGC-20854/2020\|EPI_ISL_759410\|202 | ···················· | ···················· | ···················· | ···················· | ···················· | ···················· |
| AA | 7652 | hCoV-19/Denmark/DCGC-20856/2020\|EPI_ISL_759411\|202 | ···················· | ···················· | ···················· | ···················· | ···················· | ···················· |
| AA | 7653 | hCoV-19/Denmark/DCGC-20860/2020\|EPI_ISL_759412\|202 | ···················· | ···················· | ···················· | ···················· | ···················· | ···················· |
| AA | 7654 | hCoV-19/Denmark/DCGC-20862/2020\|EPI_ISL_759413\|202 | ···················· | ·A·················· | ···················· | ···················· | ···················· | ··A················· |
| AA | 7655 | hCoV-19/Denmark/DCGC-20863/2020\|EPI_ISL_759414\|202 | ···················· | ···················· | ···················· | ···················· | ···················· | ···················· |
| AA | 7656 | hCoV-19/Denmark/DCGC-20864/2020\|EPI_ISL_759415\|202 | ···················· | ···················· | ···················· | ···················· | ···················· | ···················· |
| AA | 7657 | hCoV-19/Denmark/DCGC-20866/2020\|EPI_ISL_759416\|202 | ···················· | ···················· | ···················· | ···················· | ···················· | ···················· |
| AA | 7658 | hCoV-19/Denmark/DCGC-20867/2020\|EPI_ISL_759417\|202 | ···················· | ···················· | ···················· | ···················· | ···················· | ···················· |
| AA | 7659 | hCoV-19/Denmark/DCGC-20868/2020\|EPI_ISL_759418\|202 | ···················· | ···················· | ···················· | ···················· | ···················· | ···················· |
| AA | 7660 | hCoV-19/Denmark/DCGC-20869/2020\|EPI_ISL_759419\|202 | ···················· | ···················· | ···················· | ···················· | ···················· | ···················· |
| AA | 7661 | hCoV-19/Denmark/DCGC-20870/2020\|EPI_ISL_759420\|202 | ···················· | ···················· | ···················· | ···················· | ···················· | ···················· |
| AA | 7662 | hCoV-19/Denmark/DCGC-20873/2020\|EPI_ISL_759421\|202 | ···················· | ···················· | ···················· | ···················· | ···················· | ···················· |
| AA | 7663 | hCoV-19/Denmark/DCGC-20877/2020\|EPI_ISL_759422\|202 | ···················· | ···················· | ···················· | ···················· | ···················· | ···················· |
| AA | 7664 | hCoV-19/Denmark/DCGC-20878/2020\|EPI_ISL_759423\|202 | ···················· | ···················· | ···················· | ···················· | ···················· | ···················· |
| AA | 7665 | hCoV-19/Denmark/DCGC-20879/2020\|EPI_ISL_759424\|202 | ···················· | ···················· | ···················· | ···················· | ···················· | ···················· |
| AA | 7666 | hCoV-19/Denmark/DCGC-20880/2020\|EPI_ISL_759425\|202 | ···················· | ···················· | ···················· | ···················· | ···················· | ···················· |
| AA | 7667 | hCoV-19/Denmark/DCGC-20882/2020\|EPI_ISL_759426\|202 | ···················· | ···················· | ···················· | ···················· | ···················· | ···················· |
| AA | 7668 | hCoV-19/Denmark/DCGC-20883/2020\|EPI_ISL_759427\|202 | ···················· | ···················· | ···················· | ···················· | ···················· | ···················· |
| AA | 7669 | hCoV-19/Denmark/DCGC-20884/2020\|EPI_ISL_759428\|202 | ···················· | ···················· | ···················· | ···················· | ···················· | ···················· |
| AA | 7670 | hCoV-19/Denmark/DCGC-20885/2020\|EPI_ISL_759429\|202 | ···················· | ···················· | ···················· | ···················· | ···················· | ···················· |
| AA | 7671 | hCoV-19/Denmark/DCGC-20887/2020\|EPI_ISL_759430\|202 | ···················· | ···················· | ···················· | ···················· | ···················· | ···················· |
| AA | 7672 | hCoV-19/Denmark/DCGC-20890/2020\|EPI_ISL_759431\|202 | ···················· | ···················· | ···················· | ···················· | ···················· | ···················· |
| AA | 7673 | hCoV-19/Denmark/DCGC-20891/2020\|EPI_ISL_759432\|202 | ···················· | ···················· | ···················· | ···················· | ···················· | ···················· |
| AA | 7674 | hCoV-19/Denmark/DCGC-20893/2020\|EPI_ISL_759433\|202 | ···················· | ···················· | ···················· | ···················· | ···················· | ···················· |
| AA | 7675 | hCoV-19/Denmark/DCGC-20894/2020\|EPI_ISL_759434\|202 | ···················· | ···················· | ···················· | ···················· | ···················· | ···················· |
| AA | 7676 | hCoV-19/Denmark/DCGC-20895/2020\|EPI_ISL_759435\|202 | ···················· | ···················· | ···················· | ···················· | ···················· | ···················· |
| AA | 7677 | hCoV-19/Denmark/DCGC-20900/2020\|EPI_ISL_759436\|202 | ···················· | ···················· | ···················· | ···················· | ···················· | ···················· |
| AA | 7678 | hCoV-19/Denmark/DCGC-20904/2020\|EPI_ISL_759437\|202 | ···················· | ···················· | ···················· | ···················· | ···················· | ···················· |
| AA | 7679 | hCoV-19/Denmark/DCGC-20906/2020\|EPI_ISL_759438\|202 | ···················· | ···················· | ···················· | ···················· | ···················· | ···················· |
| AA | 7680 | hCoV-19/Denmark/DCGC-20907/2020\|EPI_ISL_759439\|202 | ···················· | ···················· | ···················· | ···················· | ···················· | ···················· |
| AA | 7681 | hCoV-19/South Korea/KDCA0186/2020\|EPI_ISL_747419\|2 | ···················· | ·A·················· | ···················· | ···················· | ···················· | ···················· |
| AA | 7682 | hCoV-19/South Korea/KDCA0187/2020\|EPI_ISL_747420\|2 | ···················· | ···················· | ···················· | ···················· | ···················· | ···················· |
| AA | 7683 | hCoV-19/South Korea/KDCA0189/2020\|EPI_ISL_747425\|2 | ···················· | ···················· | ···················· | ···················· | ···················· | ···················· |
| AA | 7684 | hCoV-19/South Korea/KDCA0190/2020\|EPI_ISL_747426\|2 | ···················· | ···················· | ···················· | ···················· | ···················· | ···················· |
| AA | 7685 | hCoV-19/South Korea/KDCA0191/2020\|EPI_ISL_747427\|2 | ···················· | ···················· | ···················· | ···················· | ···················· | ···················· |
| AA | 7686 | hCoV-19/South Korea/KDCA0192/2020\|EPI_ISL_747428\|2 | ···················· | ···················· | ···················· | ···················· | ···················· | ···················· |
| AA | 7687 | hCoV-19/South Korea/KDCA0193/2020\|EPI_ISL_747429\|2 | ···················· | ···················· | ···················· | ···················· | ···················· | ···················· |
| AA | 7688 | hCoV-19/South Korea/KDCA0194/2020\|EPI_ISL_747430\|2 | ···················· | ···················· | ···················· | ···················· | ···················· | ···················· |
| AA | 7689 | hCoV-19/South Korea/KDCA0195/2020\|EPI_ISL_747431\|2 | ···················· | ···················· | ···················· | ···················· | ···················· | ···················· |
| AA | 7690 | hCoV-19/South Korea/KDCA0197/2020\|EPI_ISL_747432\|2 | ···················· | ···················· | ···················· | ···················· | ···················· | ···················· |
| AA | 7691 | hCoV-19/South Korea/KDCA0198/2020\|EPI_ISL_747433\|2 | ···················· | ···················· | ···················· | ···················· | ···················· | ···················· |
| AA | 7692 | hCoV-19/South Korea/KDCA0200/2020\|EPI_ISL_747435\|2 | ···················· | ···················· | ···················· | ···················· | ···················· | ···················· |
| AA | 7693 | hCoV-19/South Korea/KDCA0201/2020\|EPI_ISL_747436\|2 | ···················· | ···················· | ···················· | ···················· | ···················· | ···················· |
| AA | 7694 | hCoV-19/South Korea/KDCA0202/2020\|EPI_ISL_747437\|2 | ···················· | ···················· | ···················· | ···················· | ···················· | ···················· |
| AA | 7695 | hCoV-19/South Korea/KDCA0203/2020\|EPI_ISL_747438\|2 | ···················· | ···················· | ···················· | ···················· | ···················· | ···················· |

| | | | | | | | | |
|---|---|---|---|---|---|---|---|---|
| AA | 7696 | hCoV-19/South Korea/KDCA0204/2020\|EPI_ISL_747439\|2 | .................... | .................... | .................... | .................... | .................... | .................... |
| AA | 7697 | hCoV-19/South Korea/KDCA0205/2020\|EPI_ISL_747440\|2 | .................... | .................... | .................... | .................... | .................... | .................... |
| AA | 7698 | hCoV-19/South Korea/KDCA0206/2020\|EPI_ISL_747441\|2 | .................... | .................... | .................... | .................... | .................... | .................... |
| AA | 7699 | hCoV-19/South Korea/KDCA0207/2020\|EPI_ISL_747442\|2 | .................... | .................... | .................... | .................... | .................... | .................... |
| AA | 7700 | hCoV-19/South Korea/KDCA0208/2020\|EPI_ISL_747443\|2 | .................... | .................... | .................... | .................... | .................... | .................... |
| AA | 7701 | hCoV-19/South Korea/KDCA0213/2020\|EPI_ISL_747448\|2 | .................... | .................... | .................... | .................... | .................... | .................... |
| AA | 7702 | hCoV-19/South Korea/KDCA0214/2020\|EPI_ISL_747449\|2 | .................... | .................... | .................... | .................... | .................... | .................... |
| AA | 7703 | hCoV-19/Italy/VEN-IZSVe-20RS2160-30/2020\|EPI_ISL_7 | .................... | .................... | .................... | .................... | .................... | .................... |
| AA | 7704 | hCoV-19/Italy/VEN-IZSVe-20RS2160-31/2020\|EPI_ISL_7 | .................... | .................... | .................... | .................... | .................... | .................... |
| AA | 7705 | hCoV-19/Italy/VEN-IZSVe-20RS2160-32/2020\|EPI_ISL_7 | .................... | .................... | .................... | .................... | .................... | .................... |
| AA | 7706 | hCoV-19/Italy/VEN-IZSVe-20RS2160-33/2020\|EPI_ISL_7 | .................... | .................... | .................... | .................... | .................... | .................... |
| AA | 7707 | hCoV-19/Italy/VEN-IZSVe-20RS2160-35/2020\|EPI_ISL_7 | .................... | .................... | .................... | .................... | .................... | .................... |
| AA | 7708 | hCoV-19/Italy/VEN-IZSVe-20RS2161-11/2020\|EPI_ISL_7 | .................... | .................... | .................... | .................... | .................... | .................... |
| AA | 7709 | hCoV-19/Italy/VEN-IZSVe-20RS2161-13/2020\|EPI_ISL_7 | .................... | .................... | .................... | .................... | .................... | .................... |
| AA | 7710 | hCoV-19/Italy/VEN-IZSVe-20RS2161-14/2020\|EPI_ISL_7 | .................... | .................... | .................... | .................... | .................... | .................... |
| AA | 7711 | hCoV-19/Italy/VEN-IZSVe-20RS2161-17/2020\|EPI_ISL_7 | .................... | .................... | .................... | .................... | .....T.............. | .................... |
| AA | 7712 | hCoV-19/Italy/VEN-IZSVe-20RS2161-19/2020\|EPI_ISL_7 | .................... | .................... | .................... | .................... | .....T.............. | .................... |
| AA | 7713 | hCoV-19/Italy/VEN-IZSVe-20RS2161-20/2020\|EPI_ISL_7 | .................... | .................... | .................... | .................... | .................... | .................... |
| AA | 7714 | hCoV-19/Italy/VEN-IZSVe-20RS2163-1/2020\|EPI_ISL_74 | .................... | .................... | .................... | .................... | .................... | .................... |
| AA | 7715 | hCoV-19/Italy/VEN-IZSVe-20RS2163-2/2020\|EPI_ISL_74 | .................... | .................... | .................... | .................... | .................... | .................... |
| AA | 7716 | hCoV-19/Italy/VEN-IZSVe-20RS2163-3/2020\|EPI_ISL_74 | .................... | .................... | .................... | .................... | .................... | .................... |
| AA | 7717 | hCoV-19/Italy/VEN-IZSVe-20RS2163-4/2020\|EPI_ISL_74 | .................... | .................... | .................... | .................... | .....T.............. | .................... |
| AA | 7718 | hCoV-19/Italy/VEN-IZSVe-20RS2163-6/2020\|EPI_ISL_74 | .................... | .................... | .................... | .................... | .................... | .................... |
| AA | 7719 | hCoV-19/Italy/VEN-IZSVe-20RS2163-7/2020\|EPI_ISL_74 | .................... | .................... | .................... | .................... | .................... | .................... |
| AA | 7720 | hCoV-19/Italy/VEN-IZSVe-20RS2165-10/2020\|EPI_ISL_7 | .................... | .................... | .................... | .................... | .....T.............. | .................... |
| AA | 7721 | hCoV-19/Italy/VEN-IZSVe-20RS2165-3/2020\|EPI_ISL_74 | .................... | .................... | .................... | .................... | .................... | .................... |
| AA | 7722 | hCoV-19/Italy/VEN-IZSVe-20RS2165-5/2020\|EPI_ISL_74 | .................... | .................... | .................... | .................... | .................... | .................... |
| AA | 7723 | hCoV-19/Italy/VEN-IZSVe-20RS2165-6/2020\|EPI_ISL_74 | .................... | .................... | .................... | .................... | .................... | .................... |
| AA | 7724 | hCoV-19/Italy/VEN-IZSVe-20RS2165-9/2020\|EPI_ISL_74 | .................... | .................... | .................... | .................... | .....T.............. | .................... |
| AA | 7725 | hCoV-19/Italy/VEN-IZSVe-20RS2160-28/2020\|EPI_ISL_7 | .................... | .................... | .................... | .................... | .................... | .................... |
| AA | 7726 | hCoV-19/Italy/VEN-IZSVe-20RS2160-34/2020\|EPI_ISL_7 | .................... | .................... | .................... | .................... | .................... | .................... |
| AA | 7727 | hCoV-19/Italy/VEN-IZSVe-20RS2161-12/2020\|EPI_ISL_7 | .................... | .................... | .................... | .................... | .................... | .................... |
| AA | 7728 | hCoV-19/Italy/VEN-IZSVe-20RS2161-15/2020\|EPI_ISL_7 | .................... | .................... | .................... | .................... | .................... | .................... |
| AA | 7729 | hCoV-19/Italy/VEN-IZSVe-20RS2165-1/2020\|EPI_ISL_74 | .................... | .A.................. | .................... | .................... | .................... | .................... |
| AA | 7730 | hCoV-19/Italy/VEN-IZSVe-20RS2165-2/2020\|EPI_ISL_74 | .................... | .A.................. | .................... | .................... | .................... | .................... |
| AA | 7731 | hCoV-19/Italy/VEN-IZSVe-20RS2165-4/2020\|EPI_ISL_74 | .................... | .A.................. | .................... | .................... | .................... | .................... |
| AA | 7732 | hCoV-19/Italy/VEN-IZSVe-20RS2165-7/2020\|EPI_ISL_74 | .................... | .................... | .................... | .................... | .................... | .................... |
| AA | 7733 | hCoV-19/England/204900584/2020\|EPI_ISL_747490\|2020 | .................... | .................... | .................... | .................... | .................... | .................... |
| AA | 7734 | hCoV-19/England/204900587/2020\|EPI_ISL_747491\|2020 | .................... | .................... | .................... | .................... | .................... | .................... |
| AA | 7735 | hCoV-19/England/204900588/2020\|EPI_ISL_747492\|2020 | .................... | .................... | .................... | .................... | .................... | .................... |
| AA | 7736 | hCoV-19/England/204900592/2020\|EPI_ISL_747493\|2020 | .................... | .................... | .................... | .................... | .................... | .................... |
| AA | 7737 | hCoV-19/England/204900594/2020\|EPI_ISL_747494\|2020 | .................... | .................... | .................... | .................... | .................... | .................... |
| AA | 7738 | hCoV-19/England/204940027/2020\|EPI_ISL_747495\|2020 | .................... | .................... | .................... | .................... | .................... | .................... |
| AA | 7739 | hCoV-19/England/204940073/2020\|EPI_ISL_747496\|2020 | .................... | .................... | .................... | .................... | .................... | .................... |
| AA | 7740 | hCoV-19/England/204960184/2020\|EPI_ISL_747497\|2020 | .................... | .................... | .................... | .................... | .................... | .................... |
| AA | 7741 | hCoV-19/England/205080419/2020\|EPI_ISL_747498\|2020 | .................... | .................... | .................... | .................... | .................... | .................... |
| AA | 7742 | hCoV-19/England/205080422/2020\|EPI_ISL_747499\|2020 | .................... | .................... | .................... | .................... | .................... | .................... |
| AA | 7743 | hCoV-19/Denmark/DCGC-19088/2020\|EPI_ISL_748750\|202 | .................... | .................... | .................... | .................... | .................... | .................... |
| AA | 7744 | hCoV-19/Denmark/DCGC-19090/2020\|EPI_ISL_748751\|202 | .................... | .................... | .................... | .................... | .................... | .................... |
| AA | 7745 | hCoV-19/Denmark/DCGC-19091/2020\|EPI_ISL_748752\|202 | .................... | .................... | .................... | .................... | .................... | .................... |
| AA | 7746 | hCoV-19/Denmark/DCGC-19092/2020\|EPI_ISL_748753\|202 | .................... | .................... | .................... | .................... | .................... | .................... |
| AA | 7747 | hCoV-19/Denmark/DCGC-19093/2020\|EPI_ISL_748754\|202 | .................... | .................... | .................... | .................... | .................... | .................... |
| AA | 7748 | hCoV-19/Denmark/DCGC-19094/2020\|EPI_ISL_748755\|202 | .................... | .................... | .................... | .................... | .................... | .................... |
| AA | 7749 | hCoV-19/Denmark/DCGC-19096/2020\|EPI_ISL_748756\|202 | .................... | .................... | .................... | .................... | .................... | .................... |
| AA | 7750 | hCoV-19/Denmark/DCGC-19098/2020\|EPI_ISL_748757\|202 | .................... | .................... | .................... | .................... | .................... | .................... |
| AA | 7751 | hCoV-19/Denmark/DCGC-19106/2020\|EPI_ISL_748758\|202 | .................... | .................... | .................... | .................... | .................... | .................... |
| AA | 7752 | hCoV-19/Denmark/DCGC-19107/2020\|EPI_ISL_748759\|202 | .................... | .A.................. | .................... | .................... | .................... | .................... |
| AA | 7753 | hCoV-19/Denmark/DCGC-19108/2020\|EPI_ISL_748760\|202 | .................... | .A.................. | .................... | .................... | .................... | .................... |
| AA | 7754 | hCoV-19/Denmark/DCGC-19109/2020\|EPI_ISL_748761\|202 | .................... | .................... | .................... | .................... | .................... | .................... |
| AA | 7755 | hCoV-19/Denmark/DCGC-19111/2020\|EPI_ISL_748762\|202 | .................... | .A.................. | .................... | .................... | .................... | ..A................. |
| AA | 7756 | hCoV-19/Denmark/DCGC-19112/2020\|EPI_ISL_748763\|202 | .................... | .................... | .................... | .................... | .................... | .................... |
| AA | 7757 | hCoV-19/Denmark/DCGC-19113/2020\|EPI_ISL_748764\|202 | .................... | .................... | .................... | .................... | .................... | .................... |
| AA | 7758 | hCoV-19/Denmark/DCGC-19114/2020\|EPI_ISL_748765\|202 | .................... | .................... | .................... | .................... | .................... | .................... |
| AA | 7759 | hCoV-19/Denmark/DCGC-19120/2020\|EPI_ISL_748766\|202 | .................... | .................... | .................... | .................... | .................... | .................... |
| AA | 7760 | hCoV-19/Denmark/DCGC-19122/2020\|EPI_ISL_748767\|202 | .................... | .................... | .................... | .................... | .................... | .................... |
| AA | 7761 | hCoV-19/Denmark/DCGC-19124/2020\|EPI_ISL_748768\|202 | .................... | .................... | .................... | .................... | .................... | .................... |
| AA | 7762 | hCoV-19/Denmark/DCGC-19126/2020\|EPI_ISL_748769\|202 | .................... | .................... | .................... | .................... | .................... | .................... |
| AA | 7763 | hCoV-19/Denmark/DCGC-19129/2020\|EPI_ISL_748770\|202 | .................... | .................... | .................... | .................... | .................... | .................... |
| AA | 7764 | hCoV-19/Denmark/DCGC-19131/2020\|EPI_ISL_748771\|202 | .................... | .................... | .................... | .................... | .................... | .................... |
| AA | 7765 | hCoV-19/Denmark/DCGC-19133/2020\|EPI_ISL_748772\|202 | .................... | .................... | .................... | .................... | .................... | .................... |
| AA | 7766 | hCoV-19/Denmark/DCGC-19135/2020\|EPI_ISL_748773\|202 | .................... | .................... | .................... | .................... | .................... | .................... |
| AA | 7767 | hCoV-19/Denmark/DCGC-19136/2020\|EPI_ISL_748774\|202 | .................... | .................... | .................... | .................... | .................... | .................... |
| AA | 7768 | hCoV-19/Denmark/DCGC-19139/2020\|EPI_ISL_748775\|202 | .................... | .................... | .................... | .................... | .................... | .................... |
| AA | 7769 | hCoV-19/Denmark/DCGC-19140/2020\|EPI_ISL_748776\|202 | .................... | .................... | .................... | .................... | .................... | .................... |
| AA | 7770 | hCoV-19/Denmark/DCGC-19141/2020\|EPI_ISL_748777\|202 | .................... | .................... | .................... | .................... | .................... | .................... |
| AA | 7771 | hCoV-19/Denmark/DCGC-19142/2020\|EPI_ISL_748778\|202 | .................... | .................... | .................... | .................... | .................... | .................... |
| AA | 7772 | hCoV-19/Denmark/DCGC-19169/2020\|EPI_ISL_748790\|202 | .................... | .................... | .................... | .................... | .................... | .................... |
| AA | 7773 | hCoV-19/Denmark/DCGC-19172/2020\|EPI_ISL_748791\|202 | .................... | .................... | .................... | .................... | .................... | .................... |
| AA | 7774 | hCoV-19/Denmark/DCGC-19173/2020\|EPI_ISL_748792\|202 | .................... | .................... | .................... | .................... | .................... | .................... |
| AA | 7775 | hCoV-19/Denmark/DCGC-19174/2020\|EPI_ISL_748793\|202 | .................... | .................... | .................... | .................... | .................... | .................... |

| | | | | | | | |
|---|---|---|---|---|---|---|---|
| AA | 7776 hCoV-19/Denmark/DCGC-19175/2020\|EPI_ISL_748794\|202 | ···················· | ···················· | ····················· | ···················· | ···················· | ···················· |
| AA | 7777 hCoV-19/Denmark/DCGC-19176/2020\|EPI_ISL_748795\|202 | ···················· | ···················· | ····················· | ···················· | ···················· | ···················· |
| AA | 7778 hCoV-19/Denmark/DCGC-19178/2020\|EPI_ISL_748796\|202 | ···················· | ···················· | ····················· | ···················· | ···················· | ···················· |
| AA | 7779 hCoV-19/Denmark/DCGC-19179/2020\|EPI_ISL_748797\|202 | ···················· | ···················· | ····················· | ···················· | ···················· | ···················· |
| AA | 7780 hCoV-19/Denmark/DCGC-19183/2020\|EPI_ISL_748798\|202 | ···················· | ···················· | ····················· | ···················· | ···················· | ···················· |
| AA | 7781 hCoV-19/Denmark/DCGC-19185/2020\|EPI_ISL_748799\|202 | ···················· | ·A·················· | ····················· | ···················· | ···················· | ··A················· |
| AA | 7782 hCoV-19/USA/CA-CHLA-PLM31514880/2020\|EPI_ISL_75330 | ···················· | ·A·················· | ····················· | ···················· | ···················· | ···················· |
| AA | 7783 hCoV-19/USA/CA-CHLA-PLM59752990/2020\|EPI_ISL_75330 | ···················· | ··C················· | ····················· | ···················· | ···················· | ···················· |
| AA | 7784 hCoV-19/USA/CA-CHLA-PLM93963490/2020\|EPI_ISL_75330 | ···················· | ···················· | ····················· | ···················· | ···················· | ···················· |
| AA | 7785 hCoV-19/USA/CA-CHLA-PLM11316977/2020\|EPI_ISL_75330 | ···················· | ·A·················· | ····················· | ···················· | ···················· | ···················· |
| AA | 7786 hCoV-19/USA/CA-CHLA-PLM67055611/2020\|EPI_ISL_75330 | ···················· | ···················· | ····················· | ···················· | ···················· | ···················· |
| AA | 7787 hCoV-19/USA/CA-CHLA-PLM02703185/2020\|EPI_ISL_75330 | ···················· | ···················· | ····················· | ···················· | ···················· | ···················· |
| AA | 7788 hCoV-19/USA/CA-CHLA-PLM51850514/2020\|EPI_ISL_75330 | ···················· | ···················· | ····················· | ···················· | ···················· | ···················· |
| AA | 7789 hCoV-19/USA/WI-UW-2403/2020\|EPI_ISL_754625\|2020-12 | ···················· | ···················· | ····················· | ···················· | ···················· | ···················· |
| AA | 7791 hCoV-19/USA/WI-UW-2391/2020\|EPI_ISL_754627\|2020-10 | ···················· | ···················· | ····················· | ···················· | ···················· | ···················· |
| AA | 7792 hCoV-19/USA/WI-UW-2389/2020\|EPI_ISL_754628\|2020-10 | ···················· | ···················· | ····················· | ···················· | ···················· | ···················· |
| AA | 7793 hCoV-19/USA/WI-UW-2425/2020\|EPI_ISL_754629\|2020-12 | ···················· | ···················· | ····················· | ···················· | ···················· | ···················· |
| AA | 7794 hCoV-19/USA/WI-UW-2426/2020\|EPI_ISL_754630\|2020-12 | ···················· | ···················· | ····················· | ···················· | ···················· | ···················· |
| AA | 7795 hCoV-19/USA/WI-UW-2427/2020\|EPI_ISL_754631\|2020-12 | ···················· | ···················· | ····················· | ···················· | ···················· | ···················· |
| AA | 7796 hCoV-19/USA/WI-UW-2428/2020\|EPI_ISL_754632\|2020-12 | ···················· | ··C···A············· | ····················· | ···················· | ···················· | ···················· |
| AA | 7797 hCoV-19/USA/WI-UW-2429/2020\|EPI_ISL_754633\|2020-12 | ···················· | ···················· | ····················· | ···················· | ···················· | ···················· |
| AA | 7798 hCoV-19/USA/WI-UW-2415/2020\|EPI_ISL_754634\|2020-12 | ···················· | ···················· | ····················· | ···················· | ···················· | ···················· |
| AA | 7799 hCoV-19/Canada/ON-S1519/2020\|EPI_ISL_755900\|2020-0 | ···················· | ···················· | ····················· | ···················· | ···················· | ···················· |
| AA | 7800 hCoV-19/Canada/ON-S1529/2020\|EPI_ISL_755905\|2020-0 | ···················· | ···················· | ····················· | ···················· | ···················· | ···················· |
| AA | 7801 hCoV-19/Canada/ON-S1533/2020\|EPI_ISL_755906\|2020-0 | ···················· | ·A·················· | ····················· | ···················· | ···················· | ··A················· |
| AA | 7802 hCoV-19/Canada/ON-S1536/2020\|EPI_ISL_755909\|2020-0 | ···················· | ···················· | ····················· | ···················· | ···················· | ···················· |
| AA | 7803 hCoV-19/Canada/ON-S1537/2020\|EPI_ISL_755910\|2020-0 | ···················· | ···················· | ····················· | ···················· | ···················· | ···················· |
| AA | 7804 hCoV-19/Finland/9LS1BB7/2020\|EPI_ISL_755955\|2020-1 | ···················· | ···················· | ····················· | ···················· | ···················· | ···················· |
| AA | 7805 hCoV-19/Finland/9LS1BE5/2020\|EPI_ISL_755956\|2020-1 | ···················· | ···················· | ····················· | ···················· | ···················· | ···················· |
| AA | 7806 hCoV-19/Finland/9LS1BH9/2020\|EPI_ISL_755957\|2020-1 | ···················· | ···················· | ····················· | ···················· | ···················· | ···················· |
| AA | 7807 hCoV-19/Finland/9LS5AC9/2020\|EPI_ISL_755958\|2020-1 | ···················· | ·A·················· | ····················· | ···················· | ···················· | ··A················· |
| AA | 7808 hCoV-19/Finland/9LS5AH4/2020\|EPI_ISL_755959\|2020-1 | ···················· | ·A·················· | ····················· | ···················· | ···················· | ··A················· |
| AA | 7809 hCoV-19/Finland/9LS7BD8/2020\|EPI_ISL_755960\|2020-1 | ···················· | ·A·················· | ····················· | ···················· | ···················· | ··A················· |
| AA | 7810 hCoV-19/Finland/9LS7BE11/2020\|EPI_ISL_755961\|2020- | ···················· | ···················· | ····················· | ···················· | ···················· | ···················· |
| AA | 7811 hCoV-19/Finland/9LS8BC1/2020\|EPI_ISL_755962\|2020-1 | ···················· | ·A·················· | ····················· | ···················· | ···················· | ··A················· |
| AA | 7812 hCoV-19/Finland/9LS8BF5/2020\|EPI_ISL_755963\|2020-1 | ···················· | ···················· | ····················· | ···················· | ···················· | ···················· |
| AA | 7813 hCoV-19/Finland/9LS9AF1/2020\|EPI_ISL_755964\|2020-1 | ···················· | ·A·················· | ····················· | ···················· | ···················· | ··A················· |
| AA | 7814 hCoV-19/Finland/9LS9AF5/2020\|EPI_ISL_755965\|2020-1 | ···················· | ·A·················· | ····················· | ···················· | ···················· | ··A················· |
| AA | 7815 hCoV-19/Denmark/DCGC-21361/2020\|EPI_ISL_758128\|202 | ···················· | ···················· | ····················· | ···················· | ···················· | ···················· |
| AA | 7816 hCoV-19/Denmark/DCGC-21362/2020\|EPI_ISL_758129\|202 | ···················· | ···················· | ····················· | ···················· | ···················· | ···················· |
| AA | 7817 hCoV-19/Denmark/DCGC-21363/2020\|EPI_ISL_758130\|202 | ···················· | ···················· | ····················· | ···················· | ···················· | ···················· |
| AA | 7818 hCoV-19/Denmark/DCGC-21367/2020\|EPI_ISL_758131\|202 | ···················· | ···················· | ····················· | ···················· | ···················· | ···················· |
| AA | 7819 hCoV-19/Denmark/DCGC-21368/2020\|EPI_ISL_758132\|202 | ···················· | ···················· | ····················· | ···················· | ···················· | ···················· |
| AA | 7820 hCoV-19/Denmark/DCGC-21369/2020\|EPI_ISL_758133\|202 | ···················· | ···················· | ····················· | ···················· | ···················· | ···················· |
| AA | 7821 hCoV-19/Denmark/DCGC-21370/2020\|EPI_ISL_758134\|202 | ···················· | ···················· | ····················· | ···················· | ···················· | ···················· |
| AA | 7822 hCoV-19/Denmark/DCGC-21371/2020\|EPI_ISL_758135\|202 | ···················· | ···················· | ····················· | ···················· | ···················· | ···················· |
| AA | 7823 hCoV-19/Denmark/DCGC-21372/2020\|EPI_ISL_758136\|202 | ···················· | ···················· | ····················· | ···················· | ···················· | ···················· |
| AA | 7824 hCoV-19/Denmark/DCGC-21383/2020\|EPI_ISL_758137\|202 | ···················· | ···················· | ····················· | ···················· | ···················· | ···················· |
| AA | 7825 hCoV-19/Denmark/DCGC-21384/2020\|EPI_ISL_758138\|202 | ···················· | ···················· | ····················· | ···················· | ···················· | ···················· |
| AA | 7826 hCoV-19/Denmark/DCGC-21385/2020\|EPI_ISL_758139\|202 | ···················· | ···················· | ····················· | ···················· | ···················· | ···················· |
| AA | 7827 hCoV-19/Denmark/DCGC-21390/2020\|EPI_ISL_758140\|202 | ···················· | ···················· | ····················· | ···················· | ···················· | ···················· |
| AA | 7828 hCoV-19/Denmark/DCGC-21392/2020\|EPI_ISL_758141\|202 | ···················· | ···················· | ····················· | ···················· | ···················· | ···················· |
| AA | 7829 hCoV-19/Denmark/DCGC-21393/2020\|EPI_ISL_758142\|202 | ···················· | ···················· | ····················· | ···················· | ···················· | ···················· |
| AA | 7830 hCoV-19/Denmark/DCGC-21395/2020\|EPI_ISL_758143\|202 | ···················· | ···················· | ····················· | ···················· | ···················· | ···················· |
| AA | 7831 hCoV-19/Denmark/DCGC-21397/2020\|EPI_ISL_758144\|202 | ···················· | ···················· | ····················· | ···················· | ···················· | ···················· |
| AA | 7832 hCoV-19/Denmark/DCGC-21398/2020\|EPI_ISL_758145\|202 | ···················· | ···················· | ····················· | ···················· | ···················· | ···················· |
| AA | 7833 hCoV-19/Denmark/DCGC-21400/2020\|EPI_ISL_758146\|202 | ···················· | ···················· | ····················· | ···················· | ···················· | ···················· |
| AA | 7834 hCoV-19/Denmark/DCGC-21402/2020\|EPI_ISL_758147\|202 | ···················· | ···················· | ····················· | ···················· | ···················· | ···················· |
| AA | 7835 hCoV-19/Denmark/DCGC-21405/2020\|EPI_ISL_758148\|202 | ···················· | ···················· | ····················· | ···················· | ···················· | ···················· |
| AA | 7836 hCoV-19/Denmark/DCGC-21406/2020\|EPI_ISL_758149\|202 | ···················· | ···················· | ····················· | ···················· | ···················· | ···················· |
| AA | 7837 hCoV-19/Denmark/DCGC-21407/2020\|EPI_ISL_758150\|202 | ···················· | ···················· | ····················· | ···················· | ···················· | ···················· |
| AA | 7838 hCoV-19/Denmark/DCGC-21409/2020\|EPI_ISL_758151\|202 | ···················· | ···················· | ····················· | ···················· | ···················· | ···················· |
| AA | 7839 hCoV-19/Denmark/DCGC-21410/2020\|EPI_ISL_758152\|202 | ···················· | ···················· | ····················· | ···················· | ···················· | ···················· |
| AA | 7840 hCoV-19/Denmark/DCGC-21412/2020\|EPI_ISL_758153\|202 | ···················· | ···················· | ····················· | ···················· | ···················· | ···················· |
| AA | 7841 hCoV-19/Denmark/DCGC-21413/2020\|EPI_ISL_758154\|202 | ···················· | ···················· | ····················· | ···················· | ···················· | ···················· |
| AA | 7842 hCoV-19/Denmark/DCGC-21414/2020\|EPI_ISL_758155\|202 | ···················· | ···················· | ····················· | ···················· | ···················· | ···················· |
| AA | 7843 hCoV-19/Denmark/DCGC-21415/2020\|EPI_ISL_758156\|202 | ···················· | ···················· | ····················· | ···················· | ···················· | ···················· |
| AA | 7844 hCoV-19/Denmark/DCGC-21419/2020\|EPI_ISL_758157\|202 | ···················· | ···················· | ····················· | ···················· | ···················· | ···················· |
| AA | 7845 hCoV-19/Denmark/DCGC-21420/2020\|EPI_ISL_758158\|202 | ···················· | ···················· | ····················· | ···················· | ···················· | ···················· |
| AA | 7846 hCoV-19/Denmark/DCGC-21421/2020\|EPI_ISL_758159\|202 | ···················· | ···················· | ····················· | ···················· | ···················· | ···················· |
| AA | 7847 hCoV-19/Denmark/DCGC-21428/2020\|EPI_ISL_758160\|202 | ···················· | ···················· | ····················· | ···················· | ···················· | ···················· |
| AA | 7848 hCoV-19/Denmark/DCGC-21430/2020\|EPI_ISL_758161\|202 | ···················· | ···················· | ····················· | ···················· | ···················· | ···················· |
| AA | 7849 hCoV-19/Denmark/DCGC-21431/2020\|EPI_ISL_758162\|202 | ···················· | ···················· | ···T················· | ···················· | ···················· | ···················· |
| AA | 7850 hCoV-19/Denmark/DCGC-21432/2020\|EPI_ISL_758163\|202 | ···················· | ···················· | ····················· | ···················· | ···················· | ···················· |
| AA | 7851 hCoV-19/Denmark/DCGC-21434/2020\|EPI_ISL_758164\|202 | ···················· | ···················· | ····················· | ···················· | ···················· | ···················· |
| AA | 7852 hCoV-19/Denmark/DCGC-21438/2020\|EPI_ISL_758165\|202 | ···················· | ···················· | ····················· | ···················· | ···················· | ···················· |
| AA | 7853 hCoV-19/Denmark/DCGC-21440/2020\|EPI_ISL_758166\|202 | ···················· | ···················· | ····················· | ···················· | ···················· | ···················· |
| AA | 7854 hCoV-19/Denmark/DCGC-21441/2020\|EPI_ISL_758167\|202 | ···················· | ···················· | ····················· | ···················· | ···················· | ···················· |
| AA | 7855 hCoV-19/Denmark/DCGC-21442/2020\|EPI_ISL_758168\|202 | ···················· | ···················· | ····················· | ···················· | ···················· | ···················· |
| AA | 7856 hCoV-19/Denmark/DCGC-21443/2020\|EPI_ISL_758169\|202 | ···················· | ···················· | ····················· | ···················· | ···················· | ···················· |

| | | | | | | | | |
|---|---|---|---|---|---|---|---|---|
| AA | 7857 | hCoV-19/Denmark/DCGC-21445/2020\|EPI_ISL_758170\|202 | ···················· | ···················· | ···················· | ···················· | ···················· | ···················· |
| AA | 7858 | hCoV-19/Denmark/DCGC-21447/2020\|EPI_ISL_758171\|202 | ···················· | ···················· | ···················· | ···················· | ···················· | ···················· |
| AA | 7859 | hCoV-19/Denmark/DCGC-21449/2020\|EPI_ISL_758172\|202 | ···················· | ···················· | ···················· | ···················· | ···················· | ···················· |
| AA | 7860 | hCoV-19/Denmark/DCGC-21452/2020\|EPI_ISL_758173\|202 | ···················· | ···················· | ···················· | ···················· | ···················· | ···················· |
| AA | 7861 | hCoV-19/Denmark/DCGC-21453/2020\|EPI_ISL_758174\|202 | ···················· | ···················· | ···················· | ···················· | ···················· | ···················· |
| AA | 7862 | hCoV-19/Denmark/DCGC-21454/2020\|EPI_ISL_758175\|202 | ···················· | ···················· | ···················· | ···················· | ···················· | ···················· |
| AA | 7863 | hCoV-19/Denmark/DCGC-21458/2020\|EPI_ISL_758176\|202 | ···················· | ···················· | ···················· | ···················· | ···················· | ···················· |
| AA | 7864 | hCoV-19/Denmark/DCGC-21459/2020\|EPI_ISL_758177\|202 | ···················· | ···················· | ···················· | ···················· | ···················· | ···················· |
| AA | 7865 | hCoV-19/Denmark/DCGC-21460/2020\|EPI_ISL_758178\|202 | ···················· | ···················· | ···················· | ···················· | ···················· | ···················· |
| AA | 7866 | hCoV-19/Denmark/DCGC-21462/2020\|EPI_ISL_758179\|202 | ···················· | ···················· | ···················· | ···················· | ···················· | ···················· |
| AA | 7867 | hCoV-19/Denmark/DCGC-21468/2020\|EPI_ISL_758180\|202 | ···················· | ···················· | ···················· | ···················· | ···················· | ···················· |
| AA | 7868 | hCoV-19/Denmark/DCGC-21469/2020\|EPI_ISL_758181\|202 | ···················· | ···················· | ···················· | ···················· | ···················· | ···················· |
| AA | 7869 | hCoV-19/Denmark/DCGC-21471/2020\|EPI_ISL_758182\|202 | ···················· | ···················· | ···················· | ···················· | ···················· | ···················· |
| AA | 7870 | hCoV-19/Denmark/DCGC-21472/2020\|EPI_ISL_758183\|202 | ···················· | ···················· | ···················· | ···················· | ···················· | ···················· |
| AA | 7871 | hCoV-19/Denmark/DCGC-21473/2020\|EPI_ISL_758184\|202 | ···················· | ···A················ | ···················· | ···················· | ···················· | ···················· |
| AA | 7872 | hCoV-19/Denmark/DCGC-21474/2020\|EPI_ISL_758185\|202 | ···················· | ···················· | ···················· | ···················· | ···················· | ···················· |
| AA | 7873 | hCoV-19/Denmark/DCGC-21480/2020\|EPI_ISL_758186\|202 | ···················· | ···················· | ···················· | ···················· | ···················· | ···················· |
| AA | 7874 | hCoV-19/Denmark/DCGC-21484/2020\|EPI_ISL_758187\|202 | ···················· | ···················· | ···················· | ···················· | ···················· | ···················· |
| AA | 7875 | hCoV-19/Denmark/DCGC-21487/2020\|EPI_ISL_758188\|202 | ···················· | ···················· | ···················· | ···················· | ···················· | ···················· |
| AA | 7876 | hCoV-19/Denmark/DCGC-21488/2020\|EPI_ISL_758189\|202 | ···················· | ···················· | ···················· | ···················· | ···················· | ···················· |
| AA | 7877 | hCoV-19/Denmark/DCGC-20911/2020\|EPI_ISL_759440\|202 | ···················· | ···················· | ···················· | ···················· | ···················· | ···················· |
| AA | 7878 | hCoV-19/Denmark/DCGC-20912/2020\|EPI_ISL_759441\|202 | ···················· | ···················· | ···················· | ···················· | ···················· | ···················· |
| AA | 7879 | hCoV-19/Denmark/DCGC-20913/2020\|EPI_ISL_759442\|202 | ···················· | ···················· | ···················· | ···················· | ···················· | ···················· |
| AA | 7880 | hCoV-19/Denmark/DCGC-20914/2020\|EPI_ISL_759443\|202 | ···················· | ···················· | ···················· | ···················· | ···················· | ···················· |
| AA | 7881 | hCoV-19/Denmark/DCGC-20916/2020\|EPI_ISL_759444\|202 | ···················· | ···················· | ···················· | ···················· | ···················· | ···················· |
| AA | 7882 | hCoV-19/Denmark/DCGC-20917/2020\|EPI_ISL_759445\|202 | ···················· | ···················· | ···················· | ···················· | ···················· | ···················· |
| AA | 7883 | hCoV-19/Denmark/DCGC-20918/2020\|EPI_ISL_759446\|202 | ···················· | ···················· | ···················· | ···················· | ···················· | ···················· |
| AA | 7884 | hCoV-19/Denmark/DCGC-20919/2020\|EPI_ISL_759447\|202 | ···················· | ···················· | ···················· | ···················· | ···················· | ···················· |
| AA | 7885 | hCoV-19/Denmark/DCGC-20921/2020\|EPI_ISL_759448\|202 | ···················· | ···················· | ···················· | ···················· | ···················· | ···················· |
| AA | 7886 | hCoV-19/Denmark/DCGC-20923/2020\|EPI_ISL_759449\|202 | ···················· | ···················· | ···················· | ···················· | ···················· | ···················· |
| AA | 7887 | hCoV-19/Denmark/DCGC-20924/2020\|EPI_ISL_759450\|202 | ···················· | ···················· | ···················· | ···················· | ···················· | ···················· |
| AA | 7888 | hCoV-19/Denmark/DCGC-20926/2020\|EPI_ISL_759451\|202 | ···················· | ···················· | ···················· | ···················· | ···················· | ···················· |
| AA | 7889 | hCoV-19/Denmark/DCGC-20927/2020\|EPI_ISL_759452\|202 | ···················· | ···················· | ···················· | ···················· | ···················· | ···················· |
| AA | 7890 | hCoV-19/Denmark/DCGC-20930/2020\|EPI_ISL_759453\|202 | ···················· | ···················· | ···················· | ···················· | ···················· | ···················· |
| AA | 7891 | hCoV-19/Denmark/DCGC-20932/2020\|EPI_ISL_759454\|202 | ···················· | ···················· | ···················· | ···················· | ···················· | ···················· |
| AA | 7892 | hCoV-19/Denmark/DCGC-20934/2020\|EPI_ISL_759455\|202 | ···················· | ···················· | ···················· | ···················· | ···················· | ···················· |
| AA | 7893 | hCoV-19/Denmark/DCGC-20935/2020\|EPI_ISL_759456\|202 | ···················· | ···················· | ···················· | ···················· | ···················· | ···················· |
| AA | 7894 | hCoV-19/Denmark/DCGC-20936/2020\|EPI_ISL_759457\|202 | ···················· | ·A·················· | ···················· | ···················· | ···················· | ··A················· |
| AA | 7895 | hCoV-19/Denmark/DCGC-20937/2020\|EPI_ISL_759458\|202 | ···················· | ···················· | ···················· | ···················· | ···················· | ···················· |
| AA | 7896 | hCoV-19/Denmark/DCGC-20938/2020\|EPI_ISL_759459\|202 | ···················· | ···················· | ···················· | ···················· | ···················· | ···················· |
| AA | 7897 | hCoV-19/Denmark/DCGC-20939/2020\|EPI_ISL_759460\|202 | ···················· | ···················· | ···················· | ···················· | ···················· | ···················· |
| AA | 7898 | hCoV-19/Denmark/DCGC-20942/2020\|EPI_ISL_759461\|202 | ···················· | ···················· | ···················· | ···················· | ···················· | ···················· |
| AA | 7899 | hCoV-19/Denmark/DCGC-20943/2020\|EPI_ISL_759462\|202 | ···················· | ···················· | ···················· | ···················· | ···················· | ···················· |
| AA | 7900 | hCoV-19/Denmark/DCGC-20944/2020\|EPI_ISL_759463\|202 | ···················· | ···················· | ···················· | ···················· | ···················· | ···················· |
| AA | 7901 | hCoV-19/Denmark/DCGC-20945/2020\|EPI_ISL_759464\|202 | ···················· | ···················· | ···················· | ···················· | ·····T·············· | ···················· |
| AA | 7902 | hCoV-19/Denmark/DCGC-20948/2020\|EPI_ISL_759465\|202 | ···················· | ···················· | ···················· | ···················· | ···················· | ···················· |
| AA | 7903 | hCoV-19/Denmark/DCGC-20949/2020\|EPI_ISL_759466\|202 | ···················· | ···················· | ···················· | ···················· | ···················· | ···················· |
| AA | 7904 | hCoV-19/Denmark/DCGC-20950/2020\|EPI_ISL_759467\|202 | ···················· | ···················· | ···················· | ···················· | ···················· | ···················· |
| AA | 7905 | hCoV-19/Denmark/DCGC-20951/2020\|EPI_ISL_759468\|202 | ···················· | ···················· | ···················· | ···················· | ···················· | ···················· |
| AA | 7906 | hCoV-19/Denmark/DCGC-20952/2020\|EPI_ISL_759469\|202 | ···················· | ···················· | ···················· | ···················· | ···················· | ···················· |
| AA | 7907 | hCoV-19/Denmark/DCGC-20953/2020\|EPI_ISL_759470\|202 | ···················· | ···················· | ···················· | ···················· | ···················· | ···················· |
| AA | 7908 | hCoV-19/Denmark/DCGC-20955/2020\|EPI_ISL_759471\|202 | ···················· | ···················· | ···················· | ···················· | ···················· | ···················· |
| AA | 7909 | hCoV-19/Denmark/DCGC-20956/2020\|EPI_ISL_759472\|202 | ···················· | ···················· | ···················· | ···················· | ···················· | ···················· |
| AA | 7910 | hCoV-19/Denmark/DCGC-20957/2020\|EPI_ISL_759473\|202 | ···················· | ···················· | ···················· | ···················· | ···················· | ···················· |
| AA | 7911 | hCoV-19/Denmark/DCGC-20958/2020\|EPI_ISL_759474\|202 | ···················· | ···················· | ···················· | ···················· | ···················· | ···················· |
| AA | 7912 | hCoV-19/Denmark/DCGC-20959/2020\|EPI_ISL_759475\|202 | ···················· | ···················· | ···················· | ···················· | ···················· | ···················· |
| AA | 7913 | hCoV-19/Denmark/DCGC-20960/2020\|EPI_ISL_759476\|202 | ···················· | ···················· | ···················· | ···················· | ···················· | ···················· |
| AA | 7914 | hCoV-19/Denmark/DCGC-20964/2020\|EPI_ISL_759477\|202 | ···················· | ···················· | ···················· | ···················· | ···················· | ···················· |
| AA | 7915 | hCoV-19/Denmark/DCGC-20965/2020\|EPI_ISL_759478\|202 | ···················· | ···················· | ···················· | ···················· | ···················· | ···················· |
| AA | 7916 | hCoV-19/Denmark/DCGC-20966/2020\|EPI_ISL_759479\|202 | ···················· | ···················· | ···················· | ···················· | ···················· | ···················· |
| AA | 7917 | hCoV-19/Denmark/DCGC-20968/2020\|EPI_ISL_759480\|202 | ···················· | ···················· | ···················· | ···················· | ···················· | ···················· |
| AA | 7918 | hCoV-19/Denmark/DCGC-20970/2020\|EPI_ISL_759481\|202 | ···················· | ···················· | ···················· | ···················· | ···················· | ···················· |
| AA | 7919 | hCoV-19/Denmark/DCGC-20972/2020\|EPI_ISL_759482\|202 | ···················· | ···················· | ···················· | ···················· | ···················· | ···················· |
| AA | 7920 | hCoV-19/Denmark/DCGC-20974/2020\|EPI_ISL_759483\|202 | ···················· | ···················· | ···················· | ···················· | ···················· | ···················· |
| AA | 7921 | hCoV-19/Denmark/DCGC-20975/2020\|EPI_ISL_759484\|202 | ···················· | ···················· | ···················· | ···················· | ···················· | ···················· |
| AA | 7922 | hCoV-19/Denmark/DCGC-20977/2020\|EPI_ISL_759485\|202 | ···················· | ···················· | ···················· | ···················· | ···················· | ···················· |
| AA | 7923 | hCoV-19/Denmark/DCGC-20979/2020\|EPI_ISL_759486\|202 | ···················· | ···················· | ···················· | ···················· | ···················· | ···················· |
| AA | 7924 | hCoV-19/Denmark/DCGC-20980/2020\|EPI_ISL_759487\|202 | ···················· | ···················· | ···················· | ···················· | ···················· | ···················· |
| AA | 7925 | hCoV-19/Denmark/DCGC-20982/2020\|EPI_ISL_759488\|202 | ···················· | ···················· | ···················· | ···················· | ···················· | ···················· |
| AA | 7926 | hCoV-19/Denmark/DCGC-20983/2020\|EPI_ISL_759489\|202 | ···················· | ···················· | ···················· | ···················· | ···················· | ···················· |
| AA | 7927 | hCoV-19/Denmark/DCGC-20984/2020\|EPI_ISL_759490\|202 | ···················· | ···················· | ···················· | ···················· | ···················· | ···················· |
| AA | 7928 | hCoV-19/Denmark/DCGC-20989/2020\|EPI_ISL_759491\|202 | ···················· | ···················· | ···················· | ···················· | ···················· | ···················· |
| AA | 7929 | hCoV-19/Denmark/DCGC-20993/2020\|EPI_ISL_759492\|202 | ···················· | ···················· | ···················· | ···················· | ···················· | ···················· |
| AA | 7930 | hCoV-19/Denmark/DCGC-21001/2020\|EPI_ISL_759493\|202 | ···················· | ···················· | ···················· | ···················· | ···················· | ···················· |
| AA | 7931 | hCoV-19/Denmark/DCGC-21002/2020\|EPI_ISL_759494\|202 | ···················· | ···················· | ···················· | ···················· | ···················· | ···················· |
| AA | 7932 | hCoV-19/Denmark/DCGC-21006/2020\|EPI_ISL_759495\|202 | ···················· | ···················· | ···················· | ···················· | ···················· | ···················· |
| AA | 7933 | hCoV-19/Denmark/DCGC-21007/2020\|EPI_ISL_759496\|202 | ···················· | ···················· | ···················· | ···················· | ···················· | ···················· |
| AA | 7934 | hCoV-19/Denmark/DCGC-21008/2020\|EPI_ISL_759497\|202 | ···················· | ···················· | ···················· | ···················· | ···················· | ···················· |
| AA | 7935 | hCoV-19/Denmark/DCGC-21009/2020\|EPI_ISL_759498\|202 | ···················· | ···················· | ···················· | ···················· | ···················· | ···················· |
| AA | 7936 | hCoV-19/Denmark/DCGC-21010/2020\|EPI_ISL_759499\|202 | ···················· | ···················· | ···················· | ···················· | ···················· | ···················· |

| | | | | | | | |
|---|---|---|---|---|---|---|---|
| AA | 7937 hCoV-19/Denmark/DCGC-19186/2020\|EPI_ISL_748800\|202 | ···················· | ···················· | ···················· | ···················· | ···················· | ···················· |
| AA | 7938 hCoV-19/Denmark/DCGC-19187/2020\|EPI_ISL_748801\|202 | ···················· | ·A·················· | ···················· | ···················· | ···················· | ···A················ |
| AA | 7939 hCoV-19/Denmark/DCGC-19189/2020\|EPI_ISL_748802\|202 | ···················· | ···················· | ···················· | ···················· | ···················· | ···················· |
| AA | 7940 hCoV-19/Denmark/DCGC-19190/2020\|EPI_ISL_748803\|202 | ···················· | ···················· | ···················· | ···················· | ···················· | ···················· |
| AA | 7941 hCoV-19/Denmark/DCGC-19192/2020\|EPI_ISL_748804\|202 | ···················· | ···················· | ···················· | ···················· | ···················· | ···················· |
| AA | 7942 hCoV-19/Denmark/DCGC-19193/2020\|EPI_ISL_748805\|202 | ···················· | ···················· | ···················· | ···················· | ···················· | ···················· |
| AA | 7943 hCoV-19/Denmark/DCGC-19194/2020\|EPI_ISL_748806\|202 | ···················· | ···················· | ···················· | ···················· | ···················· | ···················· |
| AA | 7944 hCoV-19/Denmark/DCGC-19196/2020\|EPI_ISL_748807\|202 | ···················· | ···················· | ···················· | ···················· | ···················· | ···················· |
| AA | 7945 hCoV-19/Denmark/DCGC-19197/2020\|EPI_ISL_748808\|202 | ···················· | ···················· | ···················· | ···················· | ···················· | ···················· |
| AA | 7946 hCoV-19/Denmark/DCGC-19198/2020\|EPI_ISL_748809\|202 | ···················· | ···················· | ···················· | ···················· | ···················· | ···················· |
| AA | 7947 hCoV-19/Denmark/DCGC-19199/2020\|EPI_ISL_748810\|202 | ···················· | ···················· | ···················· | ···················· | ···················· | ···················· |
| AA | 7948 hCoV-19/Denmark/DCGC-19200/2020\|EPI_ISL_748811\|202 | ···················· | ···················· | ···················· | ···················· | ···················· | ···················· |
| AA | 7949 hCoV-19/Denmark/DCGC-19201/2020\|EPI_ISL_748812\|202 | ···················· | ···················· | ···················· | ···················· | ···················· | ···················· |
| AA | 7950 hCoV-19/Denmark/DCGC-19203/2020\|EPI_ISL_748813\|202 | ···················· | ···················· | ···················· | ···················· | ···················· | ···················· |
| AA | 7951 hCoV-19/Denmark/DCGC-19204/2020\|EPI_ISL_748814\|202 | ···················· | ···················· | ···················· | ···················· | ···················· | ···················· |
| AA | 7952 hCoV-19/Denmark/DCGC-19206/2020\|EPI_ISL_748815\|202 | ···················· | ···················· | ···················· | ···················· | ···················· | ···················· |
| AA | 7953 hCoV-19/Denmark/DCGC-19207/2020\|EPI_ISL_748816\|202 | ···················· | ···················· | ···················· | ···················· | ···················· | ···················· |
| AA | 7954 hCoV-19/Denmark/DCGC-19208/2020\|EPI_ISL_748817\|202 | ···················· | ·A·················· | ···················· | ···················· | ···················· | ···················· |
| AA | 7955 hCoV-19/Denmark/DCGC-19209/2020\|EPI_ISL_748818\|202 | ···················· | ···················· | ···················· | ···················· | ···················· | ···················· |
| AA | 7956 hCoV-19/Denmark/DCGC-19215/2020\|EPI_ISL_748819\|202 | ···················· | ···················· | ···················· | ···················· | ···················· | ···················· |
| AA | 7957 hCoV-19/Denmark/DCGC-19216/2020\|EPI_ISL_748820\|202 | ···················· | ·A·················· | ···················· | ···················· | ···················· | ···A················ |
| AA | 7958 hCoV-19/Denmark/DCGC-19218/2020\|EPI_ISL_748821\|202 | ···················· | ···················· | ···················· | ···················· | ···················· | ···················· |
| AA | 7959 hCoV-19/Denmark/DCGC-19219/2020\|EPI_ISL_748822\|202 | ···················· | ···················· | ···················· | ···················· | ···················· | ···················· |
| AA | 7960 hCoV-19/Denmark/DCGC-19221/2020\|EPI_ISL_748823\|202 | ···················· | ···················· | ···················· | ···················· | ···················· | ···················· |
| AA | 7961 hCoV-19/Denmark/DCGC-19222/2020\|EPI_ISL_748824\|202 | ···················· | ···················· | ···················· | ···················· | ···················· | ···················· |
| AA | 7962 hCoV-19/Denmark/DCGC-19223/2020\|EPI_ISL_748825\|202 | ···················· | ···················· | ···················· | ···················· | ···················· | ···················· |
| AA | 7963 hCoV-19/Denmark/DCGC-19225/2020\|EPI_ISL_748826\|202 | ···················· | ···················· | ···················· | ···················· | ···················· | ···················· |
| AA | 7964 hCoV-19/USA/CA-CHLA-PLM32149868/2020\|EPI_ISL_75330 | ···················· | ·A·················· | ···················· | ···················· | ···················· | ···················· |
| AA | 7965 hCoV-19/USA/CA-CHLA-PLM54539532/2020\|EPI_ISL_75330 | ···················· | ·A·················· | ···················· | ···················· | ···················· | ···················· |
| AA | 7966 hCoV-19/USA/CA-CHLA-PLM61652845/2020\|EPI_ISL_75330 | ···················· | ···················· | ···················· | ···················· | ···················· | ···················· |
| AA | 7967 hCoV-19/USA/CA-CHLA-PLM40756364/2020\|EPI_ISL_75331 | ···················· | ···················· | ···················· | ···················· | ···················· | ···················· |
| AA | 7968 hCoV-19/USA/CA-CHLA-PLM87213925/2020\|EPI_ISL_75331 | ···················· | ···················· | ···················· | ···················· | ···················· | ···················· |
| AA | 7969 hCoV-19/USA/CA-CHLA-PLM46102959/2020\|EPI_ISL_75331 | ···················· | ···················· | ···················· | ···················· | ···················· | ···················· |
| AA | 7970 hCoV-19/USA/CA-CHLA-PLM13206707/2020\|EPI_ISL_75331 | ···················· | ···················· | ···················· | ···················· | ···················· | ···················· |
| AA | 7971 hCoV-19/USA/CA-CHLA-PLM32086413/2020\|EPI_ISL_75331 | ···················· | ···················· | ···················· | ···················· | ···················· | ···················· |
| AA | 7972 hCoV-19/USA/CA-CHLA-PLM86899666/2020\|EPI_ISL_75331 | ···················· | ···················· | ···················· | ···················· | ···················· | ···················· |
| AA | 7973 hCoV-19/USA/CA-CHLA-PLM94151982/2020\|EPI_ISL_75331 | ···················· | ···················· | ···················· | ···················· | ···················· | ···················· |
| AA | 7974 hCoV-19/USA/CA-CHLA-PLM78502662/2020\|EPI_ISL_75331 | ···················· | ···················· | ···················· | ···················· | ···················· | ···················· |
| AA | 7975 hCoV-19/USA/CA-CHLA-PLM11778189/2020\|EPI_ISL_75331 | ···················· | ···················· | ···················· | ···················· | ···················· | ···················· |
| AA | 7976 hCoV-19/USA/CA-CHLA-PLM02775277/2020\|EPI_ISL_75331 | ···················· | ···················· | ···················· | ···················· | ···················· | ···················· |
| AA | 7977 hCoV-19/USA/CA-CHLA-PLM40082972/2020\|EPI_ISL_75332 | ···················· | ···················· | ···················· | ···················· | ···················· | ···················· |
| AA | 7978 hCoV-19/USA/CA-CHLA-PLM40270791/2020\|EPI_ISL_75332 | ···················· | ···················· | ···················· | ···················· | ···················· | ···················· |
| AA | 7979 hCoV-19/USA/CA-CHLA-PLM86833999/2020\|EPI_ISL_75332 | ···················· | ···················· | ···················· | ···················· | ···················· | ···················· |
| AA | 7980 hCoV-19/USA/CA-CHLA-PLM57176477/2020\|EPI_ISL_75332 | ···················· | ···················· | ···················· | ···················· | ···················· | ···················· |
| AA | 7981 hCoV-19/USA/CA-CHLA-PLM97759218/2020\|EPI_ISL_75332 | ···················· | ···················· | ···················· | ···················· | ···················· | ···················· |
| AA | 7982 hCoV-19/USA/CA-CHLA-PLM47014149/2020\|EPI_ISL_75332 | ···················· | ·A·················· | ···················· | ···················· | ···················· | ···················· |
| AA | 7983 hCoV-19/USA/CA-CHLA-PLM40404799/2020\|EPI_ISL_75332 | ···················· | ···················· | ···················· | ···················· | ···················· | ···················· |
| AA | 7984 hCoV-19/USA/CA-CHLA-PLM14752712/2020\|EPI_ISL_75332 | ···················· | ···················· | ···················· | ···················· | ···················· | ···················· |
| AA | 7985 hCoV-19/USA/CA-CHLA-PLM67956098/2020\|EPI_ISL_75332 | ···················· | ···················· | ···················· | ···················· | ···················· | ···················· |
| AA | 7986 hCoV-19/USA/CA-CHLA-PLM77242671/2020\|EPI_ISL_75332 | ···················· | ···················· | ···················· | ···················· | ···················· | ···················· |
| AA | 7987 hCoV-19/USA/CA-CHLA-PLM87046575/2020\|EPI_ISL_75333 | ···················· | ···················· | ···················· | ···················· | ···················· | ···················· |
| AA | 7988 hCoV-19/USA/CA-CHLA-PLM65368962/2020\|EPI_ISL_75333 | ···················· | ···················· | ···················· | ···················· | ···················· | ···················· |
| AA | 7989 hCoV-19/USA/CA-CHLA-PLM70845114/2020\|EPI_ISL_75333 | ···················· | ···················· | ···················· | ···················· | ···················· | ···················· |
| AA | 7990 hCoV-19/USA/CA-CHLA-PLM76837845/2020\|EPI_ISL_75333 | ···················· | ·A·················· | ···················· | ···················· | ···················· | ···················· |
| AA | 7991 hCoV-19/USA/CA-CHLA-PLM66341850/2020\|EPI_ISL_75333 | ···················· | ·A·················· | ···················· | ···················· | ···················· | ···················· |
| AA | 7992 hCoV-19/USA/CA-CHLA-PLM99805263/2020\|EPI_ISL_75333 | ···················· | ···················· | ···················· | ···················· | ···················· | ···················· |
| AA | 7993 hCoV-19/USA/CA-CHLA-PLM60947497/2020\|EPI_ISL_75333 | ···················· | ···················· | ···················· | ···················· | ···················· | ···················· |
| AA | 7994 hCoV-19/USA/CA-CHLA-PLM50981298/2020\|EPI_ISL_75333 | ···················· | ·A·················· | ···················· | ···················· | ···················· | ···················· |
| AA | 7995 hCoV-19/USA/CA-CHLA-PLM28814725/2020\|EPI_ISL_75333 | ···················· | ·A·················· | ···················· | ···················· | ···················· | ···················· |
| AA | 7996 hCoV-19/USA/CA-CHLA-PLM29045953/2020\|EPI_ISL_75333 | ···················· | ·A·················· | ···················· | ···················· | ···················· | ···················· |
| AA | 7997 hCoV-19/USA/CA-CHLA-PLM90269345/2020\|EPI_ISL_75334 | ···················· | ·A·················· | ···················· | ···················· | ···················· | ···················· |
| AA | 7998 hCoV-19/USA/CA-CHLA-PLM54818837/2020\|EPI_ISL_75334 | ···················· | ···················· | ···················· | ···················· | ···················· | ···················· |
| AA | 7999 hCoV-19/USA/CA-CHLA-PLM41725240/2020\|EPI_ISL_75334 | ···················· | ·A·················· | ···················· | ···················· | ···················· | ···················· |
| AA | 8000 hCoV-19/USA/CA-CHLA-PLM80628817/2020\|EPI_ISL_75334 | ···················· | ·A·················· | ···················· | ···················· | ···················· | ···················· |
| AA | 8001 hCoV-19/USA/CA-CHLA-PLM72377620/2020\|EPI_ISL_75334 | ···················· | ···················· | ···················· | ···················· | ···················· | ···················· |
| AA | 8002 hCoV-19/USA/CA-CHLA-PLM34373484/2020\|EPI_ISL_75334 | ···················· | ·A·················· | ···················· | ···················· | ···················· | ···················· |
| AA | 8003 hCoV-19/USA/CA-CHLA-PLM96797627/2020\|EPI_ISL_75334 | ···················· | ···················· | ···················· | ···················· | ···················· | ···················· |
| AA | 8004 hCoV-19/USA/CA-CHLA-PLM07026043/2020\|EPI_ISL_75334 | ···················· | ·A·················· | ···················· | ···················· | ···················· | ···················· |
| AA | 8005 hCoV-19/USA/CA-CHLA-PLM32355874/2020\|EPI_ISL_75334 | ···················· | ···················· | ···················· | ···················· | ···················· | ···················· |
| AA | 8006 hCoV-19/USA/CA-CHLA-PLM72903549/2020\|EPI_ISL_75334 | ···················· | ···················· | ···················· | ···················· | ···················· | ···················· |
| AA | 8007 hCoV-19/USA/CA-CHLA-PLM95329786/2020\|EPI_ISL_75335 | ···················· | ···················· | ···················· | ···················· | ···················· | ···················· |
| AA | 8008 hCoV-19/USA/CA-CHLA-PLM02957000/2020\|EPI_ISL_75335 | ···················· | ···················· | ···················· | ···················· | ···················· | ···················· |
| AA | 8009 hCoV-19/USA/CA-CHLA-PLM76323778/2020\|EPI_ISL_75335 | ···················· | ·A·················· | ···················· | ···················· | ···················· | ···················· |
| AA | 8010 hCoV-19/USA/CA-CHLA-PLM78829447/2020\|EPI_ISL_75335 | ···················· | ···················· | ···················· | ···················· | ···················· | ···················· |
| AA | 8011 hCoV-19/USA/CA-CHLA-PLM79340704/2020\|EPI_ISL_75335 | ···················· | ···················· | ···················· | ···················· | ···················· | ···················· |
| AA | 8012 hCoV-19/USA/CA-CHLA-PLM07058707/2020\|EPI_ISL_75335 | ···················· | ···················· | ···················· | ···················· | ···················· | ···················· |
| AA | 8013 hCoV-19/USA/CA-CHLA-PLM78078504/2020\|EPI_ISL_75335 | ···················· | ···················· | ···················· | ···················· | ···················· | ···················· |
| AA | 8014 hCoV-19/USA/CA-CHLA-PLM16511891/2020\|EPI_ISL_75335 | ···················· | ···················· | ···················· | ···················· | ···················· | ···················· |
| AA | 8015 hCoV-19/USA/CA-CHLA-PLM26831634/2020\|EPI_ISL_75335 | ···················· | ···················· | ···················· | ···················· | ···················· | ···················· |
| AA | 8016 hCoV-19/USA/CA-CHLA-PLM12258051/2020\|EPI_ISL_75335 | ···················· | ···················· | ···················· | ···················· | ···················· | ···················· |

| | | | | | | | |
|---|---|---|---|---|---|---|---|
| AA | 8017 hCoV-19/USA/CA-CHLA-PLM35599978/2020\|EPI_ISL_75336 | ···················· | ···················· | ···················· | ···················· | ···················· | ···················· |
| AA | 8018 hCoV-19/USA/CA-CHLA-PLM03696427/2020\|EPI_ISL_75336 | ···················· | ···················· | ···················· | ···················· | ···················· | ···················· |
| AA | 8019 hCoV-19/USA/CA-CHLA-PLM67662503/2020\|EPI_ISL_75336 | ···················· | ···················· | ···················· | ···················· | ···················· | ···················· |
| AA | 8020 hCoV-19/USA/CA-CHLA-PLM04975012/2020\|EPI_ISL_75336 | ···················· | ···················· | ···················· | ···················· | ···················· | ···················· |
| AA | 8021 hCoV-19/USA/CA-CHLA-PLM07772123/2020\|EPI_ISL_75336 | ···················· | ···················· | ···················· | ···················· | ···················· | ···················· |
| AA | 8022 hCoV-19/USA/CA-CHLA-PLM92201540/2020\|EPI_ISL_75336 | ···················· | ·A·················· | ···················· | ···················· | ···················· | ···················· |
| AA | 8023 hCoV-19/USA/CA-CHLA-PLM16550109/2020\|EPI_ISL_75336 | ···················· | ···················· | ···················· | ···················· | ···················· | ···················· |
| AA | 8024 hCoV-19/USA/CA-CHLA-PLM53174364/2020\|EPI_ISL_75336 | ···················· | ···················· | ···················· | ···················· | ···················· | ···················· |
| AA | 8025 hCoV-19/USA/CA-CHLA-PLM46822648/2020\|EPI_ISL_75336 | ···················· | ···················· | ···················· | ···················· | ···················· | ···················· |
| AA | 8026 hCoV-19/USA/CA-CHLA-PLM67772232/2020\|EPI_ISL_75336 | ···················· | ···················· | ···················· | ···················· | ···················· | ···················· |
| AA | 8027 hCoV-19/USA/CA-CHLA-PLM97926928/2020\|EPI_ISL_75337 | ···················· | ·A·················· | ···················· | ···················· | ···················· | ···················· |
| AA | 8028 hCoV-19/USA/CA-CHLA-PLM21914356/2020\|EPI_ISL_75337 | ···················· | ···················· | ···················· | ···················· | ···················· | ···················· |
| AA | 8029 hCoV-19/USA/CA-CHLA-PLM81882677/2020\|EPI_ISL_75337 | ···················· | ··C················· | ···················· | ···················· | ···················· | ···················· |
| AA | 8030 hCoV-19/USA/CA-CHLA-PLM76320099/2020\|EPI_ISL_75337 | ···················· | ···················· | ···················· | ···················· | ···················· | ···················· |
| AA | 8031 hCoV-19/USA/CA-CHLA-PLM66404200/2020\|EPI_ISL_75337 | ···················· | ···················· | ···················· | ···················· | ···················· | ···················· |
| AA | 8032 hCoV-19/USA/CA-CHLA-PLM70381389/2020\|EPI_ISL_75338 | ···················· | ·A·················· | ···················· | ···················· | ···················· | ···················· |
| AA | 8033 hCoV-19/USA/CA-CHLA-PLM41302615/2020\|EPI_ISL_75338 | ···················· | ·A·················· | ···················· | ···················· | ···················· | ···················· |
| AA | 8034 hCoV-19/USA/CA-CHLA-PLM74675338/2020\|EPI_ISL_75338 | ···················· | ·A·················· | ···················· | ···················· | ···················· | ···················· |
| AA | 8035 hCoV-19/USA/CA-CHLA-PLM25366041/2020\|EPI_ISL_75338 | ···················· | ·A·················· | ···················· | ···················· | ···················· | ···················· |
| AA | 8036 hCoV-19/USA/CA-CHLA-PLM47954122/2020\|EPI_ISL_75338 | ···················· | ···················· | ···················· | ···················· | ···················· | ···················· |
| AA | 8037 hCoV-19/USA/CA-CHLA-PLM91424328/2020\|EPI_ISL_75338 | ···················· | ···················· | ···················· | ···················· | ···················· | ···················· |
| AA | 8038 hCoV-19/USA/CA-CHLA-PLM39551566/2020\|EPI_ISL_75338 | ···················· | ···················· | ···················· | ···················· | ···················· | ···················· |
| AA | 8039 hCoV-19/USA/CA-CHLA-PLM34118298/2020\|EPI_ISL_75338 | ···················· | ·A·················· | ···················· | ···················· | ···················· | ···················· |
| AA | 8040 hCoV-19/USA/CA-CHLA-PLM31438336/2020\|EPI_ISL_75338 | ···················· | ···················· | ···················· | ···················· | ···················· | ···················· |
| AA | 8041 hCoV-19/USA/CA-CHLA-PLM69146326/2020\|EPI_ISL_75339 | ···················· | ·········T·········· | ···················· | ···················· | ···················· | ···················· |
| AA | 8042 hCoV-19/USA/CA-CHLA-PLM02369650/2020\|EPI_ISL_75339 | ···················· | ···················· | ···················· | ···················· | ···················· | ···················· |
| AA | 8043 hCoV-19/USA/CA-CHLA-PLM35584098/2020\|EPI_ISL_75339 | ···················· | ·········T·········· | ···················· | ···················· | ···················· | ···················· |
| AA | 8044 hCoV-19/USA/CA-CHLA-PLM41214732/2020\|EPI_ISL_75339 | ···················· | ·········T·········· | ···················· | ···················· | ···················· | ···················· |
| AA | 8045 hCoV-19/USA/CA-CHLA-PLM27840203/2020\|EPI_ISL_75339 | ···················· | ·A·················· | ···················· | ···················· | ···················· | ···················· |
| AA | 8046 hCoV-19/USA/CA-CHLA-PLM23781419/2020\|EPI_ISL_75339 | ···················· | ·A·················· | ···················· | ···················· | ···················· | ···················· |
| AA | 8047 hCoV-19/USA/CA-CHLA-PLM20127810/2020\|EPI_ISL_75339 | ···················· | ···················· | ···················· | ···················· | ···················· | ···················· |
| AA | 8048 hCoV-19/USA/CA-CHLA-PLM16327527/2020\|EPI_ISL_75339 | ···················· | ···················· | ···················· | ···················· | ···················· | ···················· |
| AA | 8049 hCoV-19/USA/CA-CHLA-PLM38823258/2020\|EPI_ISL_75339 | ···················· | ·A·················· | ···················· | ···················· | ···················· | ··A················· |
| AA | 8050 hCoV-19/USA/CA-CHLA-PLM78687424/2020\|EPI_ISL_75339 | ···················· | ·A·················· | ···················· | ···················· | ···················· | ···················· |
| AA | 8051 hCoV-19/USA/NY-Wadsworth-278004-01/2020\|EPI_ISL_75 | ···················· | ···················· | ···················· | ···················· | ···················· | ···················· |
| AA | 8052 hCoV-19/USA/NY-Wadsworth-278033-01/2020\|EPI_ISL_75 | ···················· | ·A·················· | ···················· | ···················· | ···················· | ···················· |
| AA | 8053 hCoV-19/USA/NY-Wadsworth-278034-01/2020\|EPI_ISL_75 | ···················· | ·A·················· | ···················· | ···················· | ···················· | ···················· |
| AA | 8054 hCoV-19/USA/NY-Wadsworth-277997-01/2020\|EPI_ISL_75 | ···················· | ·A·················· | ···················· | ···················· | ···················· | ···················· |
| AA | 8055 hCoV-19/USA/NY-Wadsworth-278092-01/2020\|EPI_ISL_75 | ···················· | ·A·················· | ···················· | ···················· | ···················· | ···················· |
| AA | 8056 hCoV-19/USA/NY-Wadsworth-278015-01/2020\|EPI_ISL_75 | ···················· | ·A·················· | ···················· | ···················· | ···················· | ···················· |
| AA | 8057 hCoV-19/USA/NY-Wadsworth-277962-01/2020\|EPI_ISL_75 | ···················· | ·A·················· | ···················· | ···················· | ···················· | ···················· |
| AA | 8058 hCoV-19/USA/NY-Wadsworth-277983-01/2020\|EPI_ISL_75 | ···················· | ·A·················· | ···················· | ···················· | ···················· | ···················· |
| AA | 8059 hCoV-19/USA/NY-Wadsworth-278029-01/2020\|EPI_ISL_75 | ···················· | ·A·················· | ···················· | ···················· | ···················· | ···················· |
| AA | 8060 hCoV-19/USA/NY-Wadsworth-278030-01/2020\|EPI_ISL_75 | ···················· | ·A·················· | ···················· | ···················· | ···················· | ···················· |
| AA | 8061 hCoV-19/USA/NY-Wadsworth-278031-01/2020\|EPI_ISL_75 | ···················· | ·A·················· | ···················· | ···················· | ···················· | ···················· |
| AA | 8062 hCoV-19/USA/NY-Wadsworth-278032-01/2020\|EPI_ISL_75 | ···················· | ·A·················· | ···················· | ···················· | ···················· | ···················· |
| AA | 8063 hCoV-19/USA/NY-Wadsworth-278020-01/2020\|EPI_ISL_75 | ···················· | ·A·················· | ···················· | ···················· | ···················· | ···················· |
| AA | 8064 hCoV-19/USA/NY-Wadsworth-278021-01/2020\|EPI_ISL_75 | ···················· | ·A·················· | ···················· | ···················· | ···················· | ···················· |
| AA | 8065 hCoV-19/USA/NY-Wadsworth-278024-01/2020\|EPI_ISL_75 | ···················· | ·A·················· | ···················· | ···················· | ···················· | ···················· |
| AA | 8066 hCoV-19/USA/NY-Wadsworth-277985-01/2020\|EPI_ISL_75 | ···················· | ·A·················· | ···················· | ···················· | ···················· | ···················· |
| AA | 8067 hCoV-19/USA/NY-Wadsworth-278137-01/2020\|EPI_ISL_75 | ···················· | ·A·················· | ···················· | ···················· | ···················· | ···················· |
| AA | 8068 hCoV-19/USA/NY-Wadsworth-278126-01/2020\|EPI_ISL_75 | ···················· | ·A·················· | ···················· | ···················· | ···················· | ···················· |
| AA | 8069 hCoV-19/USA/NY-Wadsworth-278398-01/2020\|EPI_ISL_75 | ···················· | ···················· | ···················· | ···················· | ···················· | ···················· |
| AA | 8070 hCoV-19/USA/WI-UW-2411/2020\|EPI_ISL_754623\|2020-12 | ···················· | ···················· | ···················· | ···················· | ···················· | ···················· |
| AA | 8071 hCoV-19/USA/WI-UW-2417/2020\|EPI_ISL_754635\|2020-12 | ···················· | ···················· | ···················· | ···················· | ···················· | ···················· |
| AA | 8074 hCoV-19/USA/WI-UW-2416/2020\|EPI_ISL_754638\|2020-12 | ···················· | ·A·················· | ···················· | ···················· | ···················· | ···················· |
| AA | 8075 hCoV-19/USA/WI-UW-2432/2020\|EPI_ISL_754639\|2020-12 | ···················· | ···················· | ···················· | ···················· | ···················· | ···················· |
| AA | 8076 hCoV-19/USA/WI-UW-2433/2020\|EPI_ISL_754640\|2020-12 | ···················· | ·A·················· | ···················· | ···················· | ···················· | ···················· |
| AA | 8078 hCoV-19/USA/WI-UW-2443/2020\|EPI_ISL_754642\|2020-12 | ···················· | ···················· | ···················· | ···················· | ···················· | ···················· |
| AA | 8080 hCoV-19/USA/WI-UW-2396/2020\|EPI_ISL_754644\|2020-12 | ···················· | ···················· | ···················· | ···················· | ···················· | ···················· |
| AA | 8081 hCoV-19/USA/WI-UW-2435/2020\|EPI_ISL_754645\|2020-12 | ···················· | ···················· | ···················· | ···················· | ···················· | ···················· |
| AA | 8082 hCoV-19/USA/WI-UW-2439/2020\|EPI_ISL_754646\|2020-12 | ···················· | ···················· | ···················· | ···················· | ···················· | ···················· |
| AA | 8084 hCoV-19/USA/WI-UW-2392/2020\|EPI_ISL_754648\|2020-12 | ···················· | ···················· | ···················· | ···················· | ···················· | ···················· |
| AA | 8086 hCoV-19/USA/WI-UW-2431/2020\|EPI_ISL_754650\|2020-12 | ···················· | ···················· | ···················· | ···················· | ···················· | ···················· |
| AA | 8087 hCoV-19/USA/WI-UW-2436/2020\|EPI_ISL_754651\|2020-12 | ···················· | ···················· | ···················· | ···················· | ···················· | ···················· |
| AA | 8088 hCoV-19/USA/WI-UW-2440/2020\|EPI_ISL_754652\|2020-12 | ···················· | ···················· | ···················· | ···················· | ···················· | ···················· |
| AA | 8091 hCoV-19/USA/WI-UW-2430/2020\|EPI_ISL_754655\|2020-12 | ···················· | ···················· | ···················· | ···················· | ···················· | ···················· |
| AA | 8092 hCoV-19/USA/WI-UW-2434/2020\|EPI_ISL_754656\|2020-12 | ···················· | ···················· | ···················· | ···················· | ···················· | ···················· |
| AA | 8094 hCoV-19/USA/OH-UW-2400/2020\|EPI_ISL_754658\|2020-12 | ···················· | ···················· | ···················· | ···················· | ···················· | ···················· |
| AA | 8095 hCoV-19/USA/WI-UW-2404/2020\|EPI_ISL_754659\|2020-12 | ···················· | ·A·················· | ···················· | ···················· | ···················· | ···················· |
| AA | 8097 hCoV-19/USA/WI-UW-2408/2020\|EPI_ISL_754661\|2020-12 | ···················· | ···················· | ···················· | ···················· | ···················· | ···················· |
| AA | 8098 hCoV-19/USA/WI-UW-2410/2020\|EPI_ISL_754662\|2020-12 | ···················· | ···················· | ···················· | ···················· | ···················· | ···················· |
| AA | 8099 hCoV-19/USA/WI-UW-2418/2020\|EPI_ISL_754663\|2020-12 | ···················· | ···················· | ···················· | ···················· | ···················· | ···················· |
| AA | 8100 hCoV-19/USA/WI-UW-2419/2020\|EPI_ISL_754664\|2020-12 | ···················· | ···················· | ···················· | ···················· | ···················· | ···················· |
| AA | 8101 hCoV-19/USA/WI-UW-2421/2020\|EPI_ISL_754665\|2020-12 | ···················· | ···················· | ···················· | ···················· | ···················· | ···················· |
| AA | 8102 hCoV-19/USA/WI-UW-2423/2020\|EPI_ISL_754666\|2020-12 | ···················· | ···················· | ···················· | ···················· | ···················· | ···················· |
| AA | 8106 hCoV-19/USA/WI-UW-2407/2020\|EPI_ISL_754670\|2020-12 | ···················· | ···················· | ···················· | ···················· | ···················· | ···················· |
| AA | 8107 hCoV-19/USA/WI-UW-2412/2020\|EPI_ISL_754671\|2020-12 | ···················· | ···················· | ···················· | ···················· | ···················· | ···················· |
| AA | 8108 hCoV-19/USA/WI-UW-2413/2020\|EPI_ISL_754672\|2020-12 | ···················· | ···················· | ···················· | ···················· | ···················· | ···················· |
| AA | 8109 hCoV-19/USA/WI-UW-2409/2020\|EPI_ISL_754673\|2020-12 | ···················· | ···················· | ···················· | ···················· | ···················· | ···················· |

AA 8110 hCoV-19/USA/WI-UW-2414/2020|EPI_ISL_754674|2020-12 ···················· ···················· ···················· ···················· ···················· ····················
AA 8111 hCoV-19/USA/WI-UW-2422/2020|EPI_ISL_754675|2020-12 ···················· ···················· ···················· ···················· ···················· ····················
AA 8112 hCoV-19/USA/WI-UW-2388/2020|EPI_ISL_754676|2020-04 ···················· ···················· ···················· ···················· ···················· ····················
AA 8113 hCoV-19/USA/NY-Wadsworth-251059-01/2020|EPI_ISL_75 ···················· ···················· ···················· ···················· ···················· ····················
AA 8114 hCoV-19/USA/NY-Wadsworth-251060-01/2020|EPI_ISL_75 ···················· ···················· ···················· ···················· ···················· ····················
AA 8115 hCoV-19/USA/NY-Wadsworth-251061-01/2020|EPI_ISL_75 ···················· ···················· ···················· ···················· ···················· ····················
AA 8116 hCoV-19/USA/NY-Wadsworth-251063-01/2020|EPI_ISL_75 ···················· ···················· ···················· ···················· ···················· ····················
AA 8117 hCoV-19/USA/NY-Wadsworth-247857-01/2020|EPI_ISL_75 ···················· ···················· ···················· ···················· ···················· ····················
AA 8118 hCoV-19/USA/NY-Wadsworth-247856-01/2020|EPI_ISL_75 ···················· ···················· ···················· ···················· ···················· ····················
AA 8119 hCoV-19/USA/NY-Wadsworth-248128-01/2020|EPI_ISL_75 ···················· ···················· ···················· ···················· ···················· ····················
AA 8120 hCoV-19/USA/NY-Wadsworth-248124-01/2020|EPI_ISL_75 ···················· ···················· ···················· ···················· ···················· ····················
AA 8121 hCoV-19/USA/NY-Wadsworth-248120-01/2020|EPI_ISL_75 ···················· ···················· ···················· ···················· ···················· ····················
AA 8122 hCoV-19/Canada/ON-S1565/2020|EPI_ISL_755920|2020-0 ···················· ·A·················· ···················· ···················· ···················· ··A·················
AA 8123 hCoV-19/Canada/ON-S1568/2020|EPI_ISL_755921|2020-0 ···················· ···················· ···················· ···················· ···················· ····················
AA 8124 hCoV-19/Canada/ON-S1581/2020|EPI_ISL_755926|2020-0 ···················· ···················· ···················· ···················· ···················· ····················
AA 8125 hCoV-19/Canada/ON-S1582/2020|EPI_ISL_755927|2020-0 ···················· ···················· ···················· ···················· ···················· ····················
AA 8126 hCoV-19/Canada/ON-S1584/2020|EPI_ISL_755928|2020-0 ···················· ···················· ···················· ···················· ···················· ····················
AA 8127 hCoV-19/Canada/ON-S1589/2020|EPI_ISL_755930|2020-0 ···················· ···················· ···················· ···················· ···················· ····················
AA 8128 hCoV-19/Canada/ON-S1590/2020|EPI_ISL_755931|2020-0 ···················· ···················· ···················· ···················· ···················· ····················
AA 8129 hCoV-19/Canada/ON-S1591/2020|EPI_ISL_755932|2020-0 ···················· ···················· ···················· ···················· ···················· ····················
AA 8130 hCoV-19/Canada/ON-S1592/2020|EPI_ISL_755933|2020-0 ···················· ···················· ···················· ···················· ···················· ····················
AA 8131 hCoV-19/Canada/ON-S1593/2020|EPI_ISL_755934|2020-0 ···················· ···················· ···················· ···················· ···················· ····················
AA 8132 hCoV-19/Canada/ON-S1594/2020|EPI_ISL_755935|2020-0 ···················· ···················· ···················· ···················· ···················· ····················
AA 8133 hCoV-19/Canada/ON-S1595/2020|EPI_ISL_755936|2020-0 ···················· ···················· ···················· ···················· ···················· ····················
AA 8134 hCoV-19/Canada/ON-S1598/2020|EPI_ISL_755938|2020-0 ···················· ···················· ···················· ···················· ···················· ····················
AA 8135 hCoV-19/Canada/ON-S1599/2020|EPI_ISL_755939|2020-0 ···················· ···················· ···················· ···················· ···················· ····················
AA 8136 hCoV-19/USA/CA-CDPH-UC303/2020|EPI_ISL_755942|2020 ···················· ···················· ···················· ···················· ···················· ····················
AA 8137 hCoV-19/Finland/9LS11AA9/2020|EPI_ISL_755943|2020- ···················· ·A·················· ···················· ···················· ···················· ··A·················
AA 8138 hCoV-19/Finland/9LS11AC1/2020|EPI_ISL_755944|2020- ···················· ···················· ···················· ···················· ···················· ····················
AA 8139 hCoV-19/Finland/9LS11AH3/2020|EPI_ISL_755945|2020- ···················· ···················· ···················· ···················· ···················· ····················
AA 8140 hCoV-19/Finland/9LS12AG7/2020|EPI_ISL_755946|2020- ···················· ·A·················· ···················· ···················· ···················· ··A·················
AA 8141 hCoV-19/Finland/9LS13BF7/2020|EPI_ISL_755947|2020- ···················· ·A·················· ···················· ···················· ···················· ··A·················
AA 8142 hCoV-19/Finland/9LS15AF9/2020|EPI_ISL_755948|2020- ···················· ·A·················· ···················· ···················· ···················· ··A·················
AA 8143 hCoV-19/Finland/9LS16AA8/2020|EPI_ISL_755949|2020- ···················· ·A·················· ···················· ···················· ···················· ··A·················
AA 8144 hCoV-19/Finland/9LS16AA9/2020|EPI_ISL_755950|2020- ···················· ···················· ···················· ···················· ···················· ····················
AA 8145 hCoV-19/Finland/9LS16AE12/2020|EPI_ISL_755951|2020 ···················· ···················· ···················· ···················· ···················· ····················
AA 8146 hCoV-19/Finland/9LS18AA2/2020|EPI_ISL_755952|2020- ···················· ···················· ···················· ···················· ···················· ····················
AA 8147 hCoV-19/Finland/9LS18AF1/2020|EPI_ISL_755953|2020- ···················· ·A·················· ···················· ···················· ···················· ··A·················
AA 8148 hCoV-19/Finland/9LS18AH6/2020|EPI_ISL_755954|2020- ···················· ···················· ···················· ···················· ···················· ····················
AA 8149 hCoV-19/Finland/9SS15B11/2020|EPI_ISL_755966|2020- ···················· ···················· ···················· ···················· ···················· ····················
AA 8150 hCoV-19/Finland/9SS15E8/2020|EPI_ISL_755967|2020-0 ···················· ···················· ···················· ···················· ···················· ····················
AA 8151 hCoV-19/Finland/9SS20H11/2020|EPI_ISL_755968|2020- ···················· ·A·················· ···················· ···················· ···················· ··A·················
AA 8152 hCoV-19/Finland/8ES4A11/2020|EPI_ISL_755969|2020-0 ···················· ·A·················· ···················· ···················· ···················· ··A·················
AA 8153 hCoV-19/Finland/8LS10A12/2020|EPI_ISL_755970|2020- ···················· ·A·················· ···················· ···················· ···················· ··A·················
AA 8154 hCoV-19/Finland/8LS12D10/2020|EPI_ISL_755971|2020- ···················· ···················· ···················· ···················· ···················· ····················
AA 8155 hCoV-19/Finland/8LS12G9/2020|EPI_ISL_755972|2020-1 ···················· ···················· ···················· ···················· ···················· ····················
AA 8156 hCoV-19/Finland/8LS13D11/2020|EPI_ISL_755973|2020- ···················· ·A·················· ···················· ···················· ···················· ··A·················
AA 8157 hCoV-19/Finland/8LS13F1/2020|EPI_ISL_755974|2020-1 ···················· ·A·················· ···················· ···················· ···················· ··A·················
AA 8158 hCoV-19/Finland/8LS15F6/2020|EPI_ISL_755975|2020-1 ···················· ·A·················· ···················· ···················· ···················· ··A·················
AA 8159 hCoV-19/Finland/8LS18AE10/2020|EPI_ISL_755976|2020 ···················· ·A·················· ···················· ···················· ···················· ··A·················
AA 8160 hCoV-19/Finland/8LS18AF7/2020|EPI_ISL_755977|2020- ···················· ·A·················· ···················· ···················· ···················· ··A·················
AA 8161 hCoV-19/Finland/8LS18E1/2020|EPI_ISL_755978|2020-1 ···················· ·A·················· ···················· ···················· ···················· ··A·················
AA 8162 hCoV-19/Finland/8LS18H1/2020|EPI_ISL_755979|2020-1 ···················· ·A·················· ···················· ···················· ···················· ··A·················
AA 8163 hCoV-19/Finland/8LS19AD6/2020|EPI_ISL_755980|2020- ···················· ·A·················· ···················· ···················· ···················· ··A·················
AA 8164 hCoV-19/Finland/8LS19AG10/2020|EPI_ISL_755981|2020 ···················· ·A·················· ···················· ···················· ···················· ··A·················
AA 8165 hCoV-19/Finland/8LS19AG2/2020|EPI_ISL_755982|2020- ···················· ···················· ···················· ···················· ···················· ····················
AA 8166 hCoV-19/Finland/8LS19AG8/2020|EPI_ISL_755983|2020- ···················· ···················· ···················· ···················· ···················· ····················
AA 8167 hCoV-19/Finland/8LS24D7/2020|EPI_ISL_755984|2020-1 ···················· ···················· ···················· ···················· ···················· ····················
AA 8168 hCoV-19/Finland/8LS2D9/2020|EPI_ISL_755985|2020-10 ···················· ···················· ···················· ···················· ···················· ····················
AA 8169 hCoV-19/Finland/8LS2H5/2020|EPI_ISL_755986|2020-10 ···················· ·A·················· ···················· ···················· ···················· ··A·················
AA 8170 hCoV-19/Finland/8LS5E7/2020|EPI_ISL_755987|2020-10 ···················· ···················· ···················· ···················· ···················· ····················
AA 8171 hCoV-19/Finland/8LS6AF10/2020|EPI_ISL_755988|2020- ···················· ···················· ···················· ···················· ···················· ····················
AA 8172 hCoV-19/Finland/8LS6AF1/2020|EPI_ISL_755989|2020-1 ···················· ·A·················· ···················· ···················· ···················· ··A·················
AA 8173 hCoV-19/Finland/8LS6AG5/2020|EPI_ISL_755990|2020-1 ···················· ·A·················· ···················· ···················· ···················· ··A·················
AA 8174 hCoV-19/Finland/8LS9BB4/2020|EPI_ISL_755991|2020-1 ···················· ···················· ···················· ···················· ···················· ····················
AA 8175 hCoV-19/Finland/8LS9BH3/2020|EPI_ISL_755992|2020-1 ···················· ···················· ···················· ···················· ···················· ····················
AA 8176 hCoV-19/Finland/8SS13C1/2020|EPI_ISL_755993|2020-0 ···················· ···················· ···················· ···················· ···················· ····················
AA 8177 hCoV-19/Finland/8SS4C1/2020|EPI_ISL_755994|2020-09 ···················· ···················· ···················· ···················· ···················· ····················
AA 8178 hCoV-19/Finland/4LS11C5/2020|EPI_ISL_755995|2020-0 ···················· ···················· ···················· ···················· ···················· ····················
AA 8179 hCoV-19/Finland/4LS12E2/2020|EPI_ISL_755996|2020-0 ···················· ···················· ···················· ···················· ···················· ····················
AA 8180 hCoV-19/Finland/4LS19A5/2020|EPI_ISL_755997|2020-0 ···················· ···················· ···················· ···················· ···················· ····················
AA 8181 hCoV-19/Finland/4LS19F3/2020|EPI_ISL_755998|2020-0 ···················· ···················· ···················· ···················· ···················· ····················
AA 8182 hCoV-19/Finland/4LS1E1/2020|EPI_ISL_755999|2020-08 ···················· ·A·················· ···················· ···················· ···················· ··A·················
AA 8183 hCoV-19/Denmark/DCGC-21489/2020|EPI_ISL_758190|202 ···················· ···················· ···················· ···················· ···················· ····················
AA 8184 hCoV-19/Denmark/DCGC-21490/2020|EPI_ISL_758191|202 ···················· ·A·················· ···················· ···················· ···················· ····················
AA 8185 hCoV-19/Denmark/DCGC-21492/2020|EPI_ISL_758192|202 ···················· ···················· ···················· ···················· ···················· ····················
AA 8186 hCoV-19/Denmark/DCGC-21493/2020|EPI_ISL_758193|202 ···················· ···················· ···················· ···················· ···················· ····················
AA 8187 hCoV-19/Denmark/DCGC-21495/2020|EPI_ISL_758194|202 ···················· ···················· ···················· ···················· ···················· ····················
AA 8188 hCoV-19/Denmark/DCGC-21496/2020|EPI_ISL_758195|202 ···················· ···················· ···················· ···················· ···················· ····················
AA 8189 hCoV-19/Denmark/DCGC-21499/2020|EPI_ISL_758196|202 ···················· ···················· ···················· ···················· ···················· ····················

| | | | | | | | | |
|---|---|---|---|---|---|---|---|---|
| AA | 8190 | hCoV-19/Denmark/DCGC-21500/2020\|EPI_ISL_758197\|202 | ···················· | ···················· | ···················· | ···················· | ···················· | ···················· |
| AA | 8191 | hCoV-19/Denmark/DCGC-21501/2020\|EPI_ISL_758198\|202 | ···················· | ···················· | ···················· | ···················· | ···················· | ···················· |
| AA | 8192 | hCoV-19/Denmark/DCGC-21506/2020\|EPI_ISL_758199\|202 | ···················· | ···················· | ···················· | ···················· | ···················· | ···················· |
| AA | 8193 | hCoV-19/England/205080449/2020\|EPI_ISL_747500\|2020 | ···················· | ···················· | ···················· | ···················· | ···················· | ···················· |
| AA | 8194 | hCoV-19/England/205080478/2020\|EPI_ISL_747501\|2020 | ···················· | ···················· | ···················· | ···················· | ·····T·············· | ···················· |
| AA | 8195 | hCoV-19/England/205090345/2020\|EPI_ISL_747504\|2020 | ···················· | ···················· | ···················· | ···················· | ···················· | ···················· |
| AA | 8196 | hCoV-19/England/205090346/2020\|EPI_ISL_747505\|2020 | ···················· | ···················· | ···················· | ···················· | ···················· | ···················· |
| AA | 8197 | hCoV-19/England/205090348/2020\|EPI_ISL_747506\|2020 | ···················· | ···················· | ···················· | ···················· | ···················· | ···················· |
| AA | 8198 | hCoV-19/England/205090349/2020\|EPI_ISL_747507\|2020 | ···················· | ···················· | ···················· | ···················· | ···················· | ···················· |
| AA | 8199 | hCoV-19/England/205090548/2020\|EPI_ISL_747509\|2020 | ···················· | ···················· | ···················· | ···················· | ···················· | ···················· |
| AA | 8200 | hCoV-19/England/205090900/2020\|EPI_ISL_747510\|2020 | ···················· | ···················· | ···················· | ···················· | ···················· | ···················· |
| AA | 8201 | hCoV-19/England/205091069/2020\|EPI_ISL_747513\|2020 | ···················· | ···················· | ···················· | ···················· | ···················· | ···················· |
| AA | 8202 | hCoV-19/England/205091071/2020\|EPI_ISL_747514\|2020 | ···················· | ···················· | ···················· | ···················· | ···················· | ···················· |
| AA | 8203 | hCoV-19/England/205091072/2020\|EPI_ISL_747515\|2020 | ···················· | ···················· | ···················· | ···················· | ···················· | ···················· |
| AA | 8204 | hCoV-19/England/205091079/2020\|EPI_ISL_747516\|2020 | ···················· | ···················· | ···················· | ···················· | ···················· | ···················· |
| AA | 8205 | hCoV-19/England/205090256/2020\|EPI_ISL_747517\|2020 | ···················· | ···················· | ···················· | ···················· | ···················· | ···················· |
| AA | 8206 | hCoV-19/England/205090257/2020\|EPI_ISL_747518\|2020 | ···················· | ···················· | ···················· | ···················· | ···················· | ···················· |
| AA | 8207 | hCoV-19/England/205090259/2020\|EPI_ISL_747519\|2020 | ···················· | ···················· | ···················· | ···················· | ···················· | ···················· |
| AA | 8208 | hCoV-19/Denmark/DCGC-19226/2020\|EPI_ISL_748827\|202 | ···················· | ···················· | ···················· | ···················· | ···················· | ···················· |
| AA | 8209 | hCoV-19/Denmark/DCGC-19227/2020\|EPI_ISL_748828\|202 | ···················· | ···················· | ···················· | ···················· | ···················· | ···················· |
| AA | 8210 | hCoV-19/Denmark/DCGC-19229/2020\|EPI_ISL_748829\|202 | ···················· | ···················· | ···················· | ···················· | ···················· | ···················· |
| AA | 8211 | hCoV-19/Denmark/DCGC-19230/2020\|EPI_ISL_748830\|202 | ···················· | ···················· | ···················· | ···················· | ···················· | ···················· |
| AA | 8212 | hCoV-19/Denmark/DCGC-19233/2020\|EPI_ISL_748831\|202 | ···················· | ···················· | ···················· | ···················· | ···················· | ···················· |
| AA | 8213 | hCoV-19/Denmark/DCGC-19234/2020\|EPI_ISL_748832\|202 | ···················· | ···················· | ···················· | ···················· | ···················· | ···················· |
| AA | 8214 | hCoV-19/Denmark/DCGC-19235/2020\|EPI_ISL_748833\|202 | ···················· | ···················· | ···················· | ···················· | ···················· | ···················· |
| AA | 8215 | hCoV-19/Denmark/DCGC-19236/2020\|EPI_ISL_748834\|202 | ···················· | ···················· | ···················· | ···················· | ···················· | ···················· |
| AA | 8216 | hCoV-19/Denmark/DCGC-19237/2020\|EPI_ISL_748835\|202 | ···················· | ···················· | ···················· | ···················· | ···················· | ···················· |
| AA | 8217 | hCoV-19/Denmark/DCGC-19238/2020\|EPI_ISL_748836\|202 | ···················· | ···················· | ···················· | ···················· | ···················· | ···················· |
| AA | 8218 | hCoV-19/Denmark/DCGC-19239/2020\|EPI_ISL_748837\|202 | ···················· | ···················· | ···················· | ···················· | ···················· | ···················· |
| AA | 8219 | hCoV-19/Denmark/DCGC-19242/2020\|EPI_ISL_748838\|202 | ···················· | ···················· | ···················· | ···················· | ···················· | ···················· |
| AA | 8220 | hCoV-19/Denmark/DCGC-19243/2020\|EPI_ISL_748839\|202 | ···················· | ···················· | ···················· | ···················· | ···················· | ···················· |
| AA | 8221 | hCoV-19/Denmark/DCGC-19244/2020\|EPI_ISL_748840\|202 | ···················· | ···················· | ···················· | ···················· | ···················· | ···················· |
| AA | 8222 | hCoV-19/Denmark/DCGC-19245/2020\|EPI_ISL_748841\|202 | ···················· | ···················· | ···················· | ···················· | ···················· | ···················· |
| AA | 8223 | hCoV-19/Denmark/DCGC-19246/2020\|EPI_ISL_748842\|202 | ···················· | ···················· | ···················· | ···················· | ···················· | ···················· |
| AA | 8224 | hCoV-19/Denmark/DCGC-19248/2020\|EPI_ISL_748843\|202 | ···················· | ···················· | ···················· | ···················· | ···················· | ···················· |
| AA | 8225 | hCoV-19/Denmark/DCGC-19249/2020\|EPI_ISL_748844\|202 | ···················· | ···················· | ···················· | ···················· | ···················· | ···················· |
| AA | 8226 | hCoV-19/Denmark/DCGC-19250/2020\|EPI_ISL_748845\|202 | ···················· | ·A·················· | ···················· | ···················· | ···················· | ···················· |
| AA | 8227 | hCoV-19/Denmark/DCGC-19251/2020\|EPI_ISL_748846\|202 | ···················· | ···················· | ···················· | ···················· | ···················· | ···················· |
| AA | 8228 | hCoV-19/Denmark/DCGC-19252/2020\|EPI_ISL_748847\|202 | ···················· | ···················· | ···················· | ···················· | ···················· | ···················· |
| AA | 8229 | hCoV-19/Denmark/DCGC-19254/2020\|EPI_ISL_748848\|202 | ···················· | ···················· | ···················· | ···················· | ···················· | ···················· |
| AA | 8230 | hCoV-19/Denmark/DCGC-19256/2020\|EPI_ISL_748849\|202 | ···················· | ···················· | ···················· | ···················· | ···················· | ···················· |
| AA | 8231 | hCoV-19/Denmark/DCGC-19257/2020\|EPI_ISL_748850\|202 | ···················· | ···················· | ···················· | ···················· | ···················· | ···················· |
| AA | 8232 | hCoV-19/Denmark/DCGC-19259/2020\|EPI_ISL_748851\|202 | ···················· | ···················· | ···················· | ···················· | ···················· | ···················· |
| AA | 8233 | hCoV-19/Denmark/DCGC-19260/2020\|EPI_ISL_748852\|202 | ···················· | ···················· | ···················· | ···················· | ···················· | ···················· |
| AA | 8234 | hCoV-19/Denmark/DCGC-19261/2020\|EPI_ISL_748853\|202 | ···················· | ···················· | ···················· | ···················· | ···················· | ···················· |
| AA | 8235 | hCoV-19/Denmark/DCGC-19262/2020\|EPI_ISL_748854\|202 | ···················· | ···················· | ···················· | ···················· | ···················· | ···················· |
| AA | 8236 | hCoV-19/Denmark/DCGC-19263/2020\|EPI_ISL_748855\|202 | ···················· | ···················· | ···················· | ···················· | ···················· | ···················· |
| AA | 8237 | hCoV-19/Denmark/DCGC-19267/2020\|EPI_ISL_748856\|202 | ···················· | ···················· | ···················· | ···················· | ···················· | ···················· |
| AA | 8238 | hCoV-19/Denmark/DCGC-19270/2020\|EPI_ISL_748857\|202 | ···················· | ···················· | ···················· | ···················· | ···················· | ···················· |
| AA | 8239 | hCoV-19/Denmark/DCGC-19273/2020\|EPI_ISL_748858\|202 | ···················· | ···················· | ···················· | ···················· | ···················· | ···················· |
| AA | 8240 | hCoV-19/Denmark/DCGC-19275/2020\|EPI_ISL_748859\|202 | ···················· | ···················· | ···················· | ···················· | ···················· | ···················· |
| AA | 8241 | hCoV-19/Denmark/DCGC-19278/2020\|EPI_ISL_748860\|202 | ···················· | ···················· | ···················· | ···················· | ···················· | ···················· |
| AA | 8242 | hCoV-19/Denmark/DCGC-19279/2020\|EPI_ISL_748861\|202 | ···················· | ···················· | ···················· | ···················· | ···················· | ···················· |
| AA | 8243 | hCoV-19/Denmark/DCGC-19280/2020\|EPI_ISL_748862\|202 | ···················· | ···················· | ···················· | ···················· | ···················· | ···················· |
| AA | 8244 | hCoV-19/Denmark/DCGC-19281/2020\|EPI_ISL_748863\|202 | ···················· | ···················· | ···················· | ···················· | ···················· | ···················· |
| AA | 8245 | hCoV-19/Denmark/DCGC-19285/2020\|EPI_ISL_748864\|202 | ···················· | ···················· | ···················· | ···················· | ···················· | ···················· |
| AA | 8246 | hCoV-19/Denmark/DCGC-19286/2020\|EPI_ISL_748865\|202 | ···················· | ···················· | ···················· | ···················· | ···················· | ···················· |
| AA | 8247 | hCoV-19/Denmark/DCGC-19288/2020\|EPI_ISL_748866\|202 | ···················· | ···················· | ···················· | ···················· | ···················· | ···················· |
| AA | 8248 | hCoV-19/Denmark/DCGC-19293/2020\|EPI_ISL_748867\|202 | ···················· | ···················· | ···················· | ···················· | ···················· | ···················· |
| AA | 8249 | hCoV-19/Denmark/DCGC-19295/2020\|EPI_ISL_748868\|202 | ···················· | ···················· | ···················· | ···················· | ···················· | ···················· |
| AA | 8250 | hCoV-19/Denmark/DCGC-19298/2020\|EPI_ISL_748869\|202 | ···················· | ···················· | ···················· | ···················· | ···················· | ···················· |
| AA | 8251 | hCoV-19/Denmark/DCGC-19300/2020\|EPI_ISL_748870\|202 | ···················· | ···················· | ···················· | ···················· | ···················· | ···················· |
| AA | 8252 | hCoV-19/Denmark/DCGC-19302/2020\|EPI_ISL_748871\|202 | ···················· | ···················· | ···················· | ···················· | ···················· | ···················· |
| AA | 8253 | hCoV-19/Denmark/DCGC-19303/2020\|EPI_ISL_748872\|202 | ···················· | ···················· | ···················· | ···················· | ···················· | ···················· |
| AA | 8254 | hCoV-19/Denmark/DCGC-19304/2020\|EPI_ISL_748873\|202 | ···················· | ···················· | ···················· | ···················· | ···················· | ···················· |
| AA | 8255 | hCoV-19/Denmark/DCGC-19307/2020\|EPI_ISL_748874\|202 | ···················· | ···················· | ···················· | ···················· | ···················· | ···················· |
| AA | 8256 | hCoV-19/Denmark/DCGC-19310/2020\|EPI_ISL_748875\|202 | ···················· | ···················· | ···················· | ···················· | ···················· | ···················· |
| AA | 8257 | hCoV-19/Denmark/DCGC-19313/2020\|EPI_ISL_748876\|202 | ···················· | ···················· | ···················· | ···················· | ···················· | ···················· |
| AA | 8258 | hCoV-19/Denmark/DCGC-19315/2020\|EPI_ISL_748877\|202 | ···················· | ···················· | ···················· | ···················· | ···················· | ···················· |
| AA | 8259 | hCoV-19/Denmark/DCGC-19316/2020\|EPI_ISL_748878\|202 | ···················· | ···················· | ···················· | ···················· | ···················· | ···················· |
| AA | 8260 | hCoV-19/Denmark/DCGC-19318/2020\|EPI_ISL_748879\|202 | ···················· | ···················· | ···················· | ···················· | ···················· | ···················· |
| AA | 8261 | hCoV-19/Denmark/DCGC-19323/2020\|EPI_ISL_748880\|202 | ···················· | ···················· | ···················· | ···················· | ···················· | ···················· |
| AA | 8262 | hCoV-19/Denmark/DCGC-19324/2020\|EPI_ISL_748881\|202 | ···················· | ·A·················· | ···················· | ···················· | ···················· | ···················· |
| AA | 8263 | hCoV-19/Denmark/DCGC-19325/2020\|EPI_ISL_748882\|202 | ···················· | ···················· | ···················· | ···················· | ···················· | ···················· |
| AA | 8264 | hCoV-19/Denmark/DCGC-19326/2020\|EPI_ISL_748883\|202 | ···················· | ···················· | ···················· | ···················· | ···················· | ···················· |
| AA | 8265 | hCoV-19/Denmark/DCGC-19327/2020\|EPI_ISL_748884\|202 | ···················· | ···················· | ···················· | ···················· | ···················· | ···················· |
| AA | 8266 | hCoV-19/Denmark/DCGC-19328/2020\|EPI_ISL_748885\|202 | ···················· | ···················· | ···················· | ···················· | ···················· | ···················· |
| AA | 8267 | hCoV-19/Denmark/DCGC-19329/2020\|EPI_ISL_748886\|202 | ···················· | ···················· | ···················· | ···················· | ···················· | ···················· |
| AA | 8268 | hCoV-19/Denmark/DCGC-19331/2020\|EPI_ISL_748887\|202 | ···················· | ···················· | ···················· | ···················· | ···················· | ···················· |
| AA | 8269 | hCoV-19/Denmark/DCGC-19332/2020\|EPI_ISL_748888\|202 | ···················· | ···················· | ···················· | ···················· | ···················· | ···················· |

| | | | | | | | |
|---|---|---|---|---|---|---|---|
| AA | 8270 hCoV-19/Denmark/DCGC-19335/2020\|EPI_ISL_748889\|202 | .................... | .................... | .................... | .................... | .................... | .................... |
| AA | 8271 hCoV-19/Denmark/DCGC-19336/2020\|EPI_ISL_748890\|202 | .................... | .................... | .................... | .................... | .................... | .................... |
| AA | 8272 hCoV-19/Denmark/DCGC-19337/2020\|EPI_ISL_748891\|202 | .................... | .................... | .................... | .................... | .................... | .................... |
| AA | 8273 hCoV-19/Denmark/DCGC-19339/2020\|EPI_ISL_748892\|202 | .................... | .................... | .................... | .................... | .................... | .................... |
| AA | 8274 hCoV-19/Denmark/DCGC-19340/2020\|EPI_ISL_748893\|202 | .................... | .................... | .................... | .................... | .................... | .................... |
| AA | 8275 hCoV-19/Denmark/DCGC-19341/2020\|EPI_ISL_748894\|202 | .................... | .................... | .................... | .................... | .................... | .................... |
| AA | 8276 hCoV-19/Denmark/DCGC-19343/2020\|EPI_ISL_748895\|202 | .................... | .................... | .................... | .................... | .................... | .................... |
| AA | 8277 hCoV-19/Denmark/DCGC-19346/2020\|EPI_ISL_748896\|202 | .................... | .................... | .................... | .................... | .................... | .................... |
| AA | 8278 hCoV-19/Denmark/DCGC-19347/2020\|EPI_ISL_748897\|202 | .................... | .................... | .................... | .................... | .................... | .................... |
| AA | 8279 hCoV-19/Denmark/DCGC-19348/2020\|EPI_ISL_748898\|202 | .................... | .................... | .................... | .................... | .................... | .................... |
| AA | 8280 hCoV-19/Denmark/DCGC-19349/2020\|EPI_ISL_748899\|202 | .................... | .................... | .................... | .................... | .................... | .................... |
| AA | 8281 hCoV-19/USA/CA-CHLA-PLM51321793/2020\|EPI_ISL_75337 | .................... | .................... | .................... | .................... | .................... | .................... |
| AA | 8282 hCoV-19/USA/CA-CHLA-PLM34254851/2020\|EPI_ISL_75337 | .................... | .A.................. | .................... | .................... | .................... | .................... |
| AA | 8283 hCoV-19/USA/CA-CHLA-PLM95751496/2020\|EPI_ISL_75337 | .................... | .A.................. | .................... | .................... | .................... | .................... |
| AA | 8284 hCoV-19/USA/CA-CHLA-PLM18995854/2020\|EPI_ISL_75337 | .................... | .A.................. | .................... | .................... | .................... | .................... |
| AA | 8285 hCoV-19/USA/NY-Wadsworth-248117-01/2020\|EPI_ISL_75 | .................... | .................... | .................... | .................... | .................... | .................... |
| AA | 8286 hCoV-19/Denmark/DCGC-21507/2020\|EPI_ISL_758200\|202 | .................... | .................... | .................... | .................... | .................... | .................... |
| AA | 8287 hCoV-19/Denmark/DCGC-21510/2020\|EPI_ISL_758201\|202 | .................... | .................... | .................... | .................... | .................... | .................... |
| AA | 8288 hCoV-19/Denmark/DCGC-21511/2020\|EPI_ISL_758202\|202 | .................... | .................... | .................... | .................... | .................... | .................... |
| AA | 8289 hCoV-19/Denmark/DCGC-21512/2020\|EPI_ISL_758203\|202 | .................... | .................... | .................... | .................... | .................... | .................... |
| AA | 8290 hCoV-19/Denmark/DCGC-21513/2020\|EPI_ISL_758204\|202 | .................... | .................... | .................... | .................... | .................... | .................... |
| AA | 8291 hCoV-19/Denmark/DCGC-21514/2020\|EPI_ISL_758205\|202 | .................... | .................... | .................... | .................... | .................... | .................... |
| AA | 8292 hCoV-19/Denmark/DCGC-21515/2020\|EPI_ISL_758206\|202 | .................... | .................... | .................... | .................... | .................... | .................... |
| AA | 8293 hCoV-19/Denmark/DCGC-21516/2020\|EPI_ISL_758207\|202 | .................... | .................... | .................... | .................... | .................... | .................... |
| AA | 8294 hCoV-19/Denmark/DCGC-21521/2020\|EPI_ISL_758208\|202 | .................... | .................... | .................... | .................... | .................... | .................... |
| AA | 8295 hCoV-19/Denmark/DCGC-21522/2020\|EPI_ISL_758209\|202 | .................... | .................... | .................... | .................... | .................... | .................... |
| AA | 8296 hCoV-19/Denmark/DCGC-21523/2020\|EPI_ISL_758210\|202 | .................... | .................... | .................... | .................... | .................... | .................... |
| AA | 8297 hCoV-19/Denmark/DCGC-21524/2020\|EPI_ISL_758211\|202 | .................... | .................... | .................... | .................... | .................... | .................... |
| AA | 8298 hCoV-19/Denmark/DCGC-21526/2020\|EPI_ISL_758212\|202 | .................... | .................... | .................... | .................... | .................... | .................... |
| AA | 8299 hCoV-19/Denmark/DCGC-21528/2020\|EPI_ISL_758213\|202 | .................... | .................... | .................... | .................... | .................... | .................... |
| AA | 8300 hCoV-19/Denmark/DCGC-21529/2020\|EPI_ISL_758214\|202 | .................... | .................... | .................... | .................... | .................... | .................... |
| AA | 8301 hCoV-19/Denmark/DCGC-21531/2020\|EPI_ISL_758215\|202 | .................... | .................... | .................... | .................... | .................... | .................... |
| AA | 8302 hCoV-19/Denmark/DCGC-21532/2020\|EPI_ISL_758216\|202 | .................... | .................... | .................... | .................... | .................... | .................... |
| AA | 8303 hCoV-19/Denmark/DCGC-21534/2020\|EPI_ISL_758217\|202 | .................... | .................... | .................... | .................... | .................... | .................... |
| AA | 8304 hCoV-19/Denmark/DCGC-21537/2020\|EPI_ISL_758218\|202 | .................... | .................... | .................... | .................... | .................... | .................... |
| AA | 8305 hCoV-19/Denmark/DCGC-21539/2020\|EPI_ISL_758219\|202 | .................... | .................... | .................... | .................... | .................... | .................... |
| AA | 8306 hCoV-19/Denmark/DCGC-21541/2020\|EPI_ISL_758220\|202 | .................... | .................... | .................... | .................... | .................... | .................... |
| AA | 8307 hCoV-19/Denmark/DCGC-21542/2020\|EPI_ISL_758221\|202 | .................... | .................... | .................... | .................... | .................... | .................... |
| AA | 8308 hCoV-19/Denmark/DCGC-21543/2020\|EPI_ISL_758222\|202 | .................... | .................... | .................... | .................... | .................... | .................... |
| AA | 8309 hCoV-19/Denmark/DCGC-21544/2020\|EPI_ISL_758223\|202 | .................... | .................... | .................... | .................... | .................... | .................... |
| AA | 8310 hCoV-19/Denmark/DCGC-21546/2020\|EPI_ISL_758224\|202 | .................... | .................... | .................... | .................... | .................... | .................... |
| AA | 8311 hCoV-19/Denmark/DCGC-21547/2020\|EPI_ISL_758225\|202 | .................... | .................... | .................... | .................... | .................... | .................... |
| AA | 8312 hCoV-19/Denmark/DCGC-21548/2020\|EPI_ISL_758226\|202 | .................... | .................... | .................... | .................... | .................... | .................... |
| AA | 8313 hCoV-19/Denmark/DCGC-21549/2020\|EPI_ISL_758227\|202 | .................... | .................... | .................... | .................... | .................... | .................... |
| AA | 8314 hCoV-19/Denmark/DCGC-21550/2020\|EPI_ISL_758228\|202 | .................... | .................... | .................... | .................... | .................... | .................... |
| AA | 8315 hCoV-19/Denmark/DCGC-21551/2020\|EPI_ISL_758229\|202 | .................... | .................... | .................... | .................... | .................... | .................... |
| AA | 8316 hCoV-19/Denmark/DCGC-21552/2020\|EPI_ISL_758230\|202 | .................... | .................... | .................... | .................... | .................... | .................... |
| AA | 8317 hCoV-19/Denmark/DCGC-21554/2020\|EPI_ISL_758231\|202 | .................... | .................... | .................... | .................... | .................... | .................... |
| AA | 8318 hCoV-19/Denmark/DCGC-21555/2020\|EPI_ISL_758232\|202 | .................... | .................... | .................... | .................... | .................... | .................... |
| AA | 8319 hCoV-19/Denmark/DCGC-21556/2020\|EPI_ISL_758233\|202 | .................... | .................... | .................... | .................... | .................... | .................... |
| AA | 8320 hCoV-19/Denmark/DCGC-21557/2020\|EPI_ISL_758234\|202 | .................... | .................... | .................... | .................... | .................... | .................... |
| AA | 8321 hCoV-19/Denmark/DCGC-21560/2020\|EPI_ISL_758235\|202 | .................... | .................... | .................... | .................... | .................... | .................... |
| AA | 8322 hCoV-19/Denmark/DCGC-21561/2020\|EPI_ISL_758236\|202 | .................... | .................... | .................... | .................... | .................... | .................... |
| AA | 8323 hCoV-19/Denmark/DCGC-21562/2020\|EPI_ISL_758237\|202 | .................... | .................... | .................... | .................... | .................... | .................... |
| AA | 8324 hCoV-19/Denmark/DCGC-21563/2020\|EPI_ISL_758238\|202 | .................... | .................... | .................... | .................... | .................... | .................... |
| AA | 8325 hCoV-19/Denmark/DCGC-21566/2020\|EPI_ISL_758239\|202 | .................... | .................... | .................... | .................... | .................... | .................... |
| AA | 8326 hCoV-19/Denmark/DCGC-21574/2020\|EPI_ISL_758240\|202 | .................... | .................... | .................... | .................... | .................... | .................... |
| AA | 8327 hCoV-19/Denmark/DCGC-21575/2020\|EPI_ISL_758241\|202 | .................... | .................... | .................... | .................... | .................... | .................... |
| AA | 8328 hCoV-19/Denmark/DCGC-21576/2020\|EPI_ISL_758242\|202 | .................... | .................... | .................... | .................... | .................... | .................... |
| AA | 8329 hCoV-19/Denmark/DCGC-21577/2020\|EPI_ISL_758243\|202 | .................... | .................... | .................... | .................... | .................... | .................... |
| AA | 8330 hCoV-19/Denmark/DCGC-21578/2020\|EPI_ISL_758244\|202 | .................... | .................... | .................... | .................... | .................... | .................... |
| AA | 8331 hCoV-19/Denmark/DCGC-21579/2020\|EPI_ISL_758245\|202 | .................... | .................... | .................... | .................... | .................... | .................... |
| AA | 8332 hCoV-19/Denmark/DCGC-21580/2020\|EPI_ISL_758246\|202 | .................... | .................... | .................... | .................... | .................... | .................... |
| AA | 8333 hCoV-19/Denmark/DCGC-21582/2020\|EPI_ISL_758247\|202 | .................... | .................... | .................... | .................... | .................... | .................... |
| AA | 8334 hCoV-19/Denmark/DCGC-21583/2020\|EPI_ISL_758248\|202 | .................... | .................... | .................... | .................... | .................... | .................... |
| AA | 8335 hCoV-19/Denmark/DCGC-21584/2020\|EPI_ISL_758249\|202 | .................... | .................... | .................... | .................... | .................... | .................... |
| AA | 8336 hCoV-19/Denmark/DCGC-21601/2020\|EPI_ISL_758257\|202 | .................... | .A.................. | .................... | .................... | .................... | ..A................. |
| AA | 8337 hCoV-19/Denmark/DCGC-21606/2020\|EPI_ISL_758259\|202 | .................... | .................... | .................... | .................... | .................... | .................... |
| AA | 8338 hCoV-19/Denmark/DCGC-21625/2020\|EPI_ISL_758270\|202 | .................... | .................... | .................... | .................... | .................... | .................... |
| AA | 8339 hCoV-19/Denmark/DCGC-21626/2020\|EPI_ISL_758271\|202 | .................... | .................... | .................... | .................... | .................... | .................... |
| AA | 8340 hCoV-19/Denmark/DCGC-21627/2020\|EPI_ISL_758272\|202 | .................... | .................... | .................... | .................... | .................... | .................... |
| AA | 8341 hCoV-19/Denmark/DCGC-21629/2020\|EPI_ISL_758273\|202 | .................... | .................... | .................... | .................... | .................... | .................... |
| AA | 8342 hCoV-19/Denmark/DCGC-21630/2020\|EPI_ISL_758274\|202 | .................... | .................... | .................... | .................... | .................... | .................... |
| AA | 8343 hCoV-19/Denmark/DCGC-21631/2020\|EPI_ISL_758275\|202 | .................... | .................... | .................... | .................... | .................... | .................... |
| AA | 8344 hCoV-19/Denmark/DCGC-21633/2020\|EPI_ISL_758276\|202 | .................... | .A.................. | .................... | .................... | .................... | ..A................. |
| AA | 8345 hCoV-19/Denmark/DCGC-21636/2020\|EPI_ISL_758277\|202 | .................... | .................... | .................... | .................... | .................... | .................... |
| AA | 8346 hCoV-19/Denmark/DCGC-21637/2020\|EPI_ISL_758278\|202 | .................... | .................... | .................... | .................... | .................... | .................... |
| AA | 8347 hCoV-19/Denmark/DCGC-21640/2020\|EPI_ISL_758279\|202 | .................... | .................... | .................... | .................... | .................... | .................... |
| AA | 8348 hCoV-19/Denmark/DCGC-21641/2020\|EPI_ISL_758280\|202 | .................... | .................... | .................... | .................... | .................... | .................... |
| AA | 8349 hCoV-19/Denmark/DCGC-21642/2020\|EPI_ISL_758281\|202 | .................... | .................... | .................... | .................... | .................... | .................... |

| | | | | | | | | |
|---|---|---|---|---|---|---|---|---|
| AA | 8350 | hCoV-19/Denmark/DCGC-21643/2020\|EPI_ISL_758282\|202 | ···················· | ···················· | ····················· | ···················· | ···················· | ···················· |
| AA | 8351 | hCoV-19/Denmark/DCGC-21646/2020\|EPI_ISL_758283\|202 | ···················· | ···················· | ····················· | ···················· | ···················· | ···················· |
| AA | 8352 | hCoV-19/Denmark/DCGC-21648/2020\|EPI_ISL_758284\|202 | ···················· | ···················· | ····················· | ···················· | ···················· | ···················· |
| AA | 8353 | hCoV-19/Denmark/DCGC-21649/2020\|EPI_ISL_758285\|202 | ···················· | ···················· | ····················· | ···················· | ···················· | ···················· |
| AA | 8354 | hCoV-19/Denmark/DCGC-21650/2020\|EPI_ISL_758286\|202 | ···················· | ···················· | ····················· | ···················· | ···················· | ···················· |
| AA | 8355 | hCoV-19/Denmark/DCGC-21651/2020\|EPI_ISL_758287\|202 | ···················· | ···················· | ····················· | ···················· | ···················· | ···················· |
| AA | 8356 | hCoV-19/Denmark/DCGC-21652/2020\|EPI_ISL_758288\|202 | ···················· | ···················· | ····················· | ···················· | ···················· | ···················· |
| AA | 8357 | hCoV-19/Denmark/DCGC-21653/2020\|EPI_ISL_758289\|202 | ···················· | ···················· | ····················· | ···················· | ···················· | ···················· |
| AA | 8358 | hCoV-19/Denmark/DCGC-21656/2020\|EPI_ISL_758290\|202 | ···················· | ···················· | ····················· | ···················· | ···················· | ···················· |
| AA | 8359 | hCoV-19/Denmark/DCGC-21658/2020\|EPI_ISL_758291\|202 | ···················· | ···················· | ····················· | ···················· | ···················· | ···················· |
| AA | 8360 | hCoV-19/Denmark/DCGC-21659/2020\|EPI_ISL_758292\|202 | ···················· | ···················· | ····················· | ···················· | ···················· | ···················· |
| AA | 8361 | hCoV-19/Denmark/DCGC-21660/2020\|EPI_ISL_758293\|202 | ···················· | ···················· | ····················· | ···················· | ···················· | ···················· |
| AA | 8362 | hCoV-19/Denmark/DCGC-21662/2020\|EPI_ISL_758294\|202 | ···················· | ···················· | ····················· | ···················· | ···················· | ···················· |
| AA | 8363 | hCoV-19/Denmark/DCGC-21663/2020\|EPI_ISL_758295\|202 | ···················· | ···················· | ····················· | ···················· | ···················· | ···················· |
| AA | 8364 | hCoV-19/Denmark/DCGC-21664/2020\|EPI_ISL_758296\|202 | ···················· | ···················· | ····················· | ···················· | ···················· | ···················· |
| AA | 8365 | hCoV-19/Denmark/DCGC-21667/2020\|EPI_ISL_758297\|202 | ···················· | ···················· | ····················· | ···················· | ···················· | ···················· |
| AA | 8366 | hCoV-19/Denmark/DCGC-21668/2020\|EPI_ISL_758298\|202 | ···················· | ···················· | ····················· | ···················· | ···················· | ···················· |
| AA | 8367 | hCoV-19/Denmark/DCGC-21670/2020\|EPI_ISL_758299\|202 | ···················· | ···················· | ····················· | ···················· | ···················· | ···················· |
| AA | 8368 | hCoV-19/Denmark/DCGC-21012/2020\|EPI_ISL_759500\|202 | ···················· | ···················· | ····················· | ···················· | ···················· | ···················· |
| AA | 8369 | hCoV-19/Denmark/DCGC-21013/2020\|EPI_ISL_759501\|202 | ···················· | ···················· | ····················· | ···················· | ···················· | ···················· |
| AA | 8370 | hCoV-19/Denmark/DCGC-21015/2020\|EPI_ISL_759502\|202 | ···················· | ···················· | ····················· | ···················· | ···················· | ···················· |
| AA | 8371 | hCoV-19/Denmark/DCGC-21016/2020\|EPI_ISL_759503\|202 | ···················· | ·A·················· | ····················· | ···················· | ···················· | ··A················· |
| AA | 8372 | hCoV-19/Denmark/DCGC-21017/2020\|EPI_ISL_759504\|202 | ···················· | ···················· | ····················· | ···················· | ···················· | ···················· |
| AA | 8373 | hCoV-19/Denmark/DCGC-21018/2020\|EPI_ISL_759505\|202 | ···················· | ···················· | ····················· | ···················· | ···················· | ···················· |
| AA | 8374 | hCoV-19/Denmark/DCGC-21019/2020\|EPI_ISL_759506\|202 | ···················· | ···················· | ····················· | ···················· | ···················· | ···················· |
| AA | 8375 | hCoV-19/Denmark/DCGC-21020/2020\|EPI_ISL_759507\|202 | ···················· | ···················· | ····················· | ···················· | ···················· | ···················· |
| AA | 8376 | hCoV-19/Denmark/DCGC-21022/2020\|EPI_ISL_759508\|202 | ···················· | ···················· | ····················· | ···················· | ···················· | ···················· |
| AA | 8377 | hCoV-19/Denmark/DCGC-21023/2020\|EPI_ISL_759509\|202 | ···················· | ···················· | ····················· | ···················· | ···················· | ···················· |
| AA | 8378 | hCoV-19/Denmark/DCGC-21024/2020\|EPI_ISL_759510\|202 | ···················· | ···················· | ····················· | ···················· | ···················· | ···················· |
| AA | 8379 | hCoV-19/Denmark/DCGC-21025/2020\|EPI_ISL_759511\|202 | ···················· | ···················· | ····················· | ···················· | ···················· | ···················· |
| AA | 8380 | hCoV-19/Denmark/DCGC-21027/2020\|EPI_ISL_759512\|202 | ···················· | ···················· | ····················· | ···················· | ···················· | ···················· |
| AA | 8381 | hCoV-19/Denmark/DCGC-21028/2020\|EPI_ISL_759513\|202 | ···················· | ···················· | ····················· | ···················· | ···················· | ···················· |
| AA | 8382 | hCoV-19/Denmark/DCGC-21029/2020\|EPI_ISL_759514\|202 | ···················· | ···················· | ····················· | ···················· | ···················· | ···················· |
| AA | 8383 | hCoV-19/Denmark/DCGC-21031/2020\|EPI_ISL_759515\|202 | ···················· | ···················· | ····················· | ···················· | ···················· | ···················· |
| AA | 8384 | hCoV-19/Denmark/DCGC-21033/2020\|EPI_ISL_759516\|202 | ···················· | ···················· | ····················· | ···················· | ···················· | ···················· |
| AA | 8385 | hCoV-19/Denmark/DCGC-21034/2020\|EPI_ISL_759517\|202 | ···················· | ···················· | ····················· | ···················· | ···················· | ···················· |
| AA | 8386 | hCoV-19/Denmark/DCGC-21036/2020\|EPI_ISL_759518\|202 | ···················· | ···················· | ····················· | ···················· | ···················· | ···················· |
| AA | 8387 | hCoV-19/Denmark/DCGC-21038/2020\|EPI_ISL_759519\|202 | ···················· | ···················· | ····················· | ···················· | ···················· | ···················· |
| AA | 8388 | hCoV-19/Denmark/DCGC-21039/2020\|EPI_ISL_759520\|202 | ···················· | ···················· | ····················· | ···················· | ···················· | ···················· |
| AA | 8389 | hCoV-19/Denmark/DCGC-21040/2020\|EPI_ISL_759521\|202 | ···················· | ···················· | ····················· | ···················· | ···················· | ···················· |
| AA | 8390 | hCoV-19/Denmark/DCGC-21041/2020\|EPI_ISL_759522\|202 | ···················· | ···················· | ····················· | ···················· | ···················· | ···················· |
| AA | 8391 | hCoV-19/Denmark/DCGC-21046/2020\|EPI_ISL_759523\|202 | ···················· | ···················· | ····················· | ···················· | ···················· | ···················· |
| AA | 8392 | hCoV-19/Denmark/DCGC-21047/2020\|EPI_ISL_759524\|202 | ···················· | ···················· | ····················· | ···················· | ···················· | ···················· |
| AA | 8393 | hCoV-19/Denmark/DCGC-21049/2020\|EPI_ISL_759525\|202 | ···················· | ···················· | ····················· | ···················· | ···················· | ···················· |
| AA | 8394 | hCoV-19/Denmark/DCGC-21050/2020\|EPI_ISL_759526\|202 | ···················· | ···················· | ····················· | ···················· | ···················· | ···················· |
| AA | 8395 | hCoV-19/Denmark/DCGC-21053/2020\|EPI_ISL_759527\|202 | ···················· | ···················· | ····················· | ···················· | ···················· | ···················· |
| AA | 8396 | hCoV-19/Denmark/DCGC-21055/2020\|EPI_ISL_759528\|202 | ···················· | ···················· | ····················· | ···················· | ···················· | ···················· |
| AA | 8397 | hCoV-19/Denmark/DCGC-21057/2020\|EPI_ISL_759529\|202 | ···················· | ···················· | ····················· | ···················· | ···················· | ···············A···· |
| AA | 8398 | hCoV-19/Denmark/DCGC-21058/2020\|EPI_ISL_759530\|202 | ···················· | ···················· | ····················· | ···················· | ···················· | ···················· |
| AA | 8399 | hCoV-19/Denmark/DCGC-21061/2020\|EPI_ISL_759531\|202 | ···················· | ···················· | ····················· | ···················· | ···················· | ···················· |
| AA | 8400 | hCoV-19/Denmark/DCGC-21063/2020\|EPI_ISL_759532\|202 | ···················· | ···················· | ····················· | ···················· | ···················· | ···················· |
| AA | 8401 | hCoV-19/Denmark/DCGC-21064/2020\|EPI_ISL_759533\|202 | ···················· | ···················· | ····················· | ···················· | ···················· | ···················· |
| AA | 8402 | hCoV-19/Denmark/DCGC-21065/2020\|EPI_ISL_759534\|202 | ···················· | ···················· | ····················· | ···················· | ···················· | ···················· |
| AA | 8403 | hCoV-19/Denmark/DCGC-21068/2020\|EPI_ISL_759535\|202 | ···················· | ···················· | ····················· | ···················· | ···················· | ···················· |
| AA | 8404 | hCoV-19/Denmark/DCGC-21071/2020\|EPI_ISL_759536\|202 | ···················· | ···················· | ····················· | ···················· | ···················· | ···················· |
| AA | 8405 | hCoV-19/Denmark/DCGC-21075/2020\|EPI_ISL_759537\|202 | ···················· | ···················· | ····················· | ···················· | ···················· | ···················· |
| AA | 8406 | hCoV-19/Denmark/DCGC-21077/2020\|EPI_ISL_759538\|202 | ···················· | ···················· | ····················· | ···················· | ···················· | ···················· |
| AA | 8407 | hCoV-19/Denmark/DCGC-21078/2020\|EPI_ISL_759539\|202 | ···················· | ···················· | ····················· | ···················· | ···················· | ···················· |
| AA | 8408 | hCoV-19/Denmark/DCGC-21079/2020\|EPI_ISL_759540\|202 | ···················· | ···················· | ····················· | ···················· | ···················· | ···················· |
| AA | 8409 | hCoV-19/Denmark/DCGC-21083/2020\|EPI_ISL_759541\|202 | ···················· | ···················· | ····················· | ···················· | ···················· | ···················· |
| AA | 8410 | hCoV-19/Denmark/DCGC-21084/2020\|EPI_ISL_759542\|202 | ···················· | ···················· | ····················· | ···················· | ···················· | ···················· |
| AA | 8411 | hCoV-19/Denmark/DCGC-21085/2020\|EPI_ISL_759543\|202 | ···················· | ···················· | ····················· | ···················· | ···················· | ···················· |
| AA | 8412 | hCoV-19/Denmark/DCGC-21086/2020\|EPI_ISL_759544\|202 | ···················· | ···················· | ····················· | ···················· | ···················· | ···················· |
| AA | 8413 | hCoV-19/Denmark/DCGC-21087/2020\|EPI_ISL_759545\|202 | ···················· | ···················· | ····················· | ···················· | ···················· | ···················· |
| AA | 8414 | hCoV-19/Denmark/DCGC-21088/2020\|EPI_ISL_759546\|202 | ···················· | ···················· | ····················· | ···················· | ···················· | ···················· |
| AA | 8415 | hCoV-19/Denmark/DCGC-21090/2020\|EPI_ISL_759547\|202 | ···················· | ···················· | ····················· | ···················· | ···················· | ···················· |
| AA | 8416 | hCoV-19/Denmark/DCGC-21091/2020\|EPI_ISL_759548\|202 | ···················· | ···················· | ····················· | ···················· | ···················· | ···················· |
| AA | 8417 | hCoV-19/Denmark/DCGC-21092/2020\|EPI_ISL_759549\|202 | ···················· | ···················· | ····················· | ···················· | ···················· | ···················· |
| AA | 8418 | hCoV-19/Denmark/DCGC-21093/2020\|EPI_ISL_759550\|202 | ···················· | ·A·················· | ····················· | ···················· | ···················· | ··A················· |
| AA | 8419 | hCoV-19/Denmark/DCGC-21095/2020\|EPI_ISL_759551\|202 | ···················· | ···················· | ····················· | ···················· | ···················· | ···················· |
| AA | 8420 | hCoV-19/Denmark/DCGC-21096/2020\|EPI_ISL_759552\|202 | ···················· | ···················· | ····················· | ···················· | ···················· | ···················· |
| AA | 8421 | hCoV-19/Denmark/DCGC-21097/2020\|EPI_ISL_759553\|202 | ···················· | ·A·················· | ····················· | ···················· | ···················· | ··A················· |
| AA | 8422 | hCoV-19/Denmark/DCGC-21098/2020\|EPI_ISL_759554\|202 | ···················· | ···················· | ····················· | ···················· | ···················· | ···················· |
| AA | 8423 | hCoV-19/Denmark/DCGC-21099/2020\|EPI_ISL_759555\|202 | ···················· | ···················· | ····················· | ···················· | ···················· | ···················· |
| AA | 8424 | hCoV-19/Denmark/DCGC-21103/2020\|EPI_ISL_759556\|202 | ···················· | ···················· | ····················· | ···················· | ···················· | ···················· |
| AA | 8425 | hCoV-19/Denmark/DCGC-21104/2020\|EPI_ISL_759557\|202 | ···················· | ···················· | ····················· | ···················· | ···················· | ···················· |
| AA | 8426 | hCoV-19/Denmark/DCGC-21106/2020\|EPI_ISL_759558\|202 | ···················· | ···················· | ····················· | ···················· | ···················· | ···················· |
| AA | 8427 | hCoV-19/Denmark/DCGC-21110/2020\|EPI_ISL_759559\|202 | ···················· | ···················· | ····················· | ···················· | ···················· | ···················· |
| AA | 8428 | hCoV-19/Denmark/DCGC-21115/2020\|EPI_ISL_759560\|202 | ···················· | ···················· | ····················· | ···················· | ···················· | ···················· |
| AA | 8429 | hCoV-19/Denmark/DCGC-21117/2020\|EPI_ISL_759561\|202 | ···················· | ···················· | ····················· | ···················· | ···················· | ···················· |

| | | | | | | | |
|---|---|---|---|---|---|---|---|
| AA | 8430 hCoV-19/Denmark/DCGC-21118/2020\|EPI_ISL_759562\|202 | ···················· | ···················· | ···················· | ···················· | ···················· | ···················· |
| AA | 8431 hCoV-19/Denmark/DCGC-21120/2020\|EPI_ISL_759563\|202 | ···················· | ···················· | ···················· | ···················· | ···················· | ···················· |
| AA | 8432 hCoV-19/Denmark/DCGC-21121/2020\|EPI_ISL_759564\|202 | ···················· | ···················· | ···················· | ···················· | ···················· | ···················· |
| AA | 8433 hCoV-19/Denmark/DCGC-21124/2020\|EPI_ISL_759565\|202 | ···················· | ···················· | ···················· | ···················· | ···················· | ···················· |
| AA | 8434 hCoV-19/Denmark/DCGC-21125/2020\|EPI_ISL_759566\|202 | ···················· | ···················· | ···················· | ···················· | ···················· | ···················· |
| AA | 8435 hCoV-19/Denmark/DCGC-21126/2020\|EPI_ISL_759567\|202 | ···················· | ···················· | ···················· | ···················· | ···················· | ···················· |
| AA | 8436 hCoV-19/Denmark/DCGC-21127/2020\|EPI_ISL_759568\|202 | ···················· | ···················· | ···················· | ···················· | ···················· | ···················· |
| AA | 8437 hCoV-19/Denmark/DCGC-21128/2020\|EPI_ISL_759569\|202 | ···················· | ···················· | ···················· | ···················· | ···················· | ···················· |
| AA | 8438 hCoV-19/Denmark/DCGC-21129/2020\|EPI_ISL_759570\|202 | ···················· | ···················· | ···················· | ···················· | ···················· | ···················· |
| AA | 8439 hCoV-19/Denmark/DCGC-21130/2020\|EPI_ISL_759571\|202 | ···················· | ···················· | ···················· | ···················· | ···················· | ···················· |
| AA | 8440 hCoV-19/Denmark/DCGC-21133/2020\|EPI_ISL_759572\|202 | ···················· | ···················· | ···················· | ···················· | ···················· | ···················· |
| AA | 8441 hCoV-19/Denmark/DCGC-21136/2020\|EPI_ISL_759573\|202 | ···················· | ···················· | ···················· | ···················· | ···················· | ···················· |
| AA | 8442 hCoV-19/Denmark/DCGC-21141/2020\|EPI_ISL_759574\|202 | ···················· | ···················· | ···················· | ···················· | ···················· | ···················· |
| AA | 8443 hCoV-19/Denmark/DCGC-21142/2020\|EPI_ISL_759575\|202 | ···················· | ···················· | ···················· | ···················· | ···················· | ···················· |
| AA | 8444 hCoV-19/Denmark/DCGC-21144/2020\|EPI_ISL_759576\|202 | ···················· | ···················· | ···················· | ···················· | ···················· | ···················· |
| AA | 8445 hCoV-19/Denmark/DCGC-21145/2020\|EPI_ISL_759577\|202 | ···················· | ···················· | ···················· | ···················· | ···················· | ···················· |
| AA | 8446 hCoV-19/Denmark/DCGC-21152/2020\|EPI_ISL_759578\|202 | ···················· | ···················· | ···················· | ···················· | ···················· | ···················· |
| AA | 8447 hCoV-19/Denmark/DCGC-21153/2020\|EPI_ISL_759579\|202 | ···················· | ···················· | ···················· | ···················· | ···················· | ···················· |
| AA | 8448 hCoV-19/Denmark/DCGC-21169/2020\|EPI_ISL_759588\|202 | ···················· | ···················· | ···················· | ···················· | ···················· | ···················· |
| AA | 8449 hCoV-19/Denmark/DCGC-19899/2020\|EPI_ISL_747612\|202 | ···················· | ···················· | ···················· | ···················· | ···················· | ···················· |
| AA | 8450 hCoV-19/Denmark/DCGC-19907/2020\|EPI_ISL_747613\|202 | ···················· | ···················· | ···················· | ···················· | ···················· | ···················· |
| AA | 8451 hCoV-19/Denmark/DCGC-19911/2020\|EPI_ISL_747614\|202 | ···················· | ···················· | ···················· | ···················· | ···················· | ···················· |
| AA | 8452 hCoV-19/Uruguay/RIV-M5/2020\|EPI_ISL_747615\|2020-05 | ···················· | ···················· | ···················· | ···················· | ···················· | ···················· |
| AA | 8453 hCoV-19/Denmark/DCGC-19938/2020\|EPI_ISL_747616\|202 | ···················· | ···················· | ···················· | ···················· | ···················· | ···················· |
| AA | 8454 hCoV-19/Denmark/DCGC-19350/2020\|EPI_ISL_748900\|202 | ···················· | ···················· | ···················· | ···················· | ···················· | ···················· |
| AA | 8455 hCoV-19/Denmark/DCGC-19352/2020\|EPI_ISL_748901\|202 | ···················· | ···················· | ···················· | ···················· | ···················· | ···················· |
| AA | 8456 hCoV-19/Denmark/DCGC-19355/2020\|EPI_ISL_748902\|202 | ···················· | ···················· | ···················· | ···················· | ···················· | ···················· |
| AA | 8457 hCoV-19/Denmark/DCGC-19356/2020\|EPI_ISL_748903\|202 | ···················· | ···················· | ···················· | ···················· | ···················· | ···················· |
| AA | 8458 hCoV-19/Denmark/DCGC-19360/2020\|EPI_ISL_748904\|202 | ···················· | ···················· | ···················· | ···················· | ···················· | ···················· |
| AA | 8459 hCoV-19/Denmark/DCGC-19361/2020\|EPI_ISL_748905\|202 | ···················· | ···················· | ···················· | ···················· | ···················· | ···················· |
| AA | 8460 hCoV-19/Denmark/DCGC-19366/2020\|EPI_ISL_748908\|202 | ···················· | ···················· | ···················· | ···················· | ···················· | ···················· |
| AA | 8461 hCoV-19/Denmark/DCGC-19368/2020\|EPI_ISL_748909\|202 | ···················· | ···················· | ···················· | ···················· | ···················· | ···················· |
| AA | 8462 hCoV-19/Denmark/DCGC-19371/2020\|EPI_ISL_748910\|202 | ···················· | ···················· | ···················· | ···················· | ···················· | ···················· |
| AA | 8463 hCoV-19/Denmark/DCGC-19372/2020\|EPI_ISL_748911\|202 | ···················· | ···················· | ···················· | ···················· | ···················· | ···················· |
| AA | 8464 hCoV-19/Denmark/DCGC-19373/2020\|EPI_ISL_748912\|202 | ···················· | ···················· | ···················· | ···················· | ···················· | ···················· |
| AA | 8465 hCoV-19/Denmark/DCGC-19374/2020\|EPI_ISL_748913\|202 | ···················· | ···················· | ···················· | ···················· | ···················· | ···················· |
| AA | 8466 hCoV-19/Denmark/DCGC-19375/2020\|EPI_ISL_748914\|202 | ···················· | ···················· | ···················· | ···················· | ···················· | ···················· |
| AA | 8467 hCoV-19/Denmark/DCGC-19376/2020\|EPI_ISL_748915\|202 | ···················· | ···················· | ···················· | ···················· | ···················· | ···················· |
| AA | 8468 hCoV-19/Denmark/DCGC-19379/2020\|EPI_ISL_748916\|202 | ···················· | ···················· | ···················· | ···················· | ···················· | ···················· |
| AA | 8469 hCoV-19/Denmark/DCGC-19381/2020\|EPI_ISL_748917\|202 | ···················· | ···················· | ···················· | ···················· | ···················· | ···················· |
| AA | 8470 hCoV-19/Denmark/DCGC-19387/2020\|EPI_ISL_748918\|202 | ···················· | ···················· | ···················· | ···················· | ···················· | ···················· |
| AA | 8471 hCoV-19/Denmark/DCGC-19389/2020\|EPI_ISL_748919\|202 | ···················· | ···················· | ···················· | ···················· | ···················· | ···················· |
| AA | 8472 hCoV-19/Denmark/DCGC-19390/2020\|EPI_ISL_748920\|202 | ···················· | ···················· | ···················· | ···················· | ···················· | ···················· |
| AA | 8473 hCoV-19/Denmark/DCGC-19392/2020\|EPI_ISL_748921\|202 | ···················· | ···················· | ···················· | ···················· | ···················· | ···················· |
| AA | 8474 hCoV-19/Denmark/DCGC-19393/2020\|EPI_ISL_748922\|202 | ···················· | ···················· | ···················· | ···················· | ···················· | ···················· |
| AA | 8475 hCoV-19/Denmark/DCGC-19394/2020\|EPI_ISL_748923\|202 | ···················· | ···················· | ···················· | ···················· | ···················· | ···················· |
| AA | 8476 hCoV-19/Denmark/DCGC-19395/2020\|EPI_ISL_748924\|202 | ···················· | ···················· | ···················· | ···················· | ···················· | ···················· |
| AA | 8477 hCoV-19/Denmark/DCGC-19398/2020\|EPI_ISL_748925\|202 | ···················· | ·A·················· | ···················· | ···················· | ···················· | ···················· |
| AA | 8478 hCoV-19/Denmark/DCGC-19399/2020\|EPI_ISL_748926\|202 | ···················· | ···················· | ···················· | ···················· | ···················· | ···················· |
| AA | 8479 hCoV-19/Denmark/DCGC-19400/2020\|EPI_ISL_748927\|202 | ···················· | ·A·················· | ···················· | ···················· | ···················· | ···················· |
| AA | 8480 hCoV-19/Denmark/DCGC-19402/2020\|EPI_ISL_748928\|202 | ···················· | ···················· | ···················· | ···················· | ···················· | ···················· |
| AA | 8481 hCoV-19/Denmark/DCGC-19404/2020\|EPI_ISL_748929\|202 | ···················· | ···················· | ···················· | ···················· | ···················· | ···················· |
| AA | 8482 hCoV-19/Denmark/DCGC-19406/2020\|EPI_ISL_748930\|202 | ···················· | ···················· | ···················· | ···················· | ···················· | ···················· |
| AA | 8483 hCoV-19/Denmark/DCGC-19410/2020\|EPI_ISL_748931\|202 | ···················· | ···················· | ···················· | ···················· | ···················· | ···················· |
| AA | 8484 hCoV-19/Denmark/DCGC-19411/2020\|EPI_ISL_748932\|202 | ···················· | ···················· | ···················· | ···················· | ···················· | ···················· |
| AA | 8485 hCoV-19/Denmark/DCGC-19412/2020\|EPI_ISL_748933\|202 | ···················· | ···················· | ···················· | ···················· | ···················· | ···················· |
| AA | 8486 hCoV-19/Denmark/DCGC-19420/2020\|EPI_ISL_748934\|202 | ···················· | ···················· | ···················· | ···················· | ···················· | ···················· |
| AA | 8487 hCoV-19/Denmark/DCGC-19423/2020\|EPI_ISL_748935\|202 | ···················· | ···················· | ···················· | ···················· | ···················· | ···················· |
| AA | 8488 hCoV-19/Denmark/DCGC-19424/2020\|EPI_ISL_748936\|202 | ···················· | ···················· | ···················· | ···················· | ···················· | ···················· |
| AA | 8489 hCoV-19/Denmark/DCGC-19426/2020\|EPI_ISL_748937\|202 | ···················· | ···················· | ···················· | ···················· | ···················· | ···················· |
| AA | 8490 hCoV-19/Denmark/DCGC-19429/2020\|EPI_ISL_748938\|202 | ···················· | ···················· | ···················· | ···················· | ···················· | ···················· |
| AA | 8491 hCoV-19/Denmark/DCGC-19430/2020\|EPI_ISL_748939\|202 | ···················· | ·A·················· | ···················· | ···················· | ···················· | ···················· |
| AA | 8492 hCoV-19/Denmark/DCGC-19431/2020\|EPI_ISL_748940\|202 | ···················· | ···················· | ···················· | ···················· | ···················· | ···················· |
| AA | 8493 hCoV-19/Denmark/DCGC-19432/2020\|EPI_ISL_748941\|202 | ···················· | ···················· | ···················· | ···················· | ···················· | ···················· |
| AA | 8494 hCoV-19/Denmark/DCGC-19433/2020\|EPI_ISL_748942\|202 | ···················· | ···················· | ···················· | ···················· | ···················· | ···················· |
| AA | 8495 hCoV-19/Denmark/DCGC-19434/2020\|EPI_ISL_748943\|202 | ···················· | ···················· | ···················· | ···················· | ···················· | ···················· |
| AA | 8496 hCoV-19/Denmark/DCGC-19435/2020\|EPI_ISL_748944\|202 | ···················· | ···················· | ···················· | ···················· | ···················· | ···················· |
| AA | 8497 hCoV-19/Denmark/DCGC-19437/2020\|EPI_ISL_748945\|202 | ···················· | ···················· | ··T················· | ···················· | ···················· | ···················· |
| AA | 8498 hCoV-19/Denmark/DCGC-19438/2020\|EPI_ISL_748946\|202 | ···················· | ·A·················· | ···················· | ···················· | ···················· | ··A················· |
| AA | 8499 hCoV-19/Denmark/DCGC-19439/2020\|EPI_ISL_748947\|202 | ···················· | ···················· | ···················· | ···················· | ···················· | ···················· |
| AA | 8500 hCoV-19/USA/CA-CHLA-PLM88317027/2020\|EPI_ISL_75340 | ···················· | ···················· | ···················· | ···················· | ···················· | ···················· |
| AA | 8501 hCoV-19/USA/CA-CHLA-PLM45839909/2020\|EPI_ISL_75340 | ···················· | ···················· | ···················· | ···················· | ···················· | ···················· |
| AA | 8502 hCoV-19/USA/CA-CHLA-PLM32737996/2020\|EPI_ISL_75340 | ···················· | ···················· | ···················· | ···················· | ···················· | ···················· |
| AA | 8503 hCoV-19/USA/CA-CHLA-PLM48677009/2020\|EPI_ISL_75340 | ···················· | ···················· | ···················· | ···················· | ···················· | ···················· |
| AA | 8504 hCoV-19/USA/CA-CHLA-PLM10942198/2020\|EPI_ISL_75340 | ···················· | ···················· | ···················· | ···················· | ···················· | ···················· |
| AA | 8505 hCoV-19/USA/CA-CHLA-PLM13300872/2020\|EPI_ISL_75340 | ···················· | ···················· | ···················· | ···················· | ···················· | ···················· |
| AA | 8506 hCoV-19/USA/CA-CHLA-PLM25383132/2020\|EPI_ISL_75340 | ···················· | ·A·················· | ···················· | ···················· | ···················· | ···················· |
| AA | 8507 hCoV-19/USA/CA-CHLA-PLM39538022/2020\|EPI_ISL_75340 | ···················· | ·A·················· | ···················· | ···················· | ···················· | ···················· |
| AA | 8508 hCoV-19/USA/CA-CHLA-PLM38887875/2020\|EPI_ISL_75340 | ···················· | ·A·················· | ···················· | ···················· | ···················· | ···················· |
| AA | 8509 hCoV-19/USA/CA-CHLA-PLM57057181/2020\|EPI_ISL_75341 | ···················· | ···················· | ···················· | ···················· | ···················· | ···················· |

```
AA    8510 hCoV-19/USA/CA-CHLA-PLM78059394/2020|EPI_ISL_75341    ····················  ····················  ····················  ····················  ····················  ····················
AA    8511 hCoV-19/USA/CA-CHLA-PLM36012316/2020|EPI_ISL_75341    ····················  ····················  ····················  ····················  ····················  ····················
AA    8512 hCoV-19/USA/CA-CHLA-PLM99900003/2020|EPI_ISL_75341    ····················  ·A··················  ····················  ····················  ····················  ····················
AA    8513 hCoV-19/USA/CA-CHLA-PLM49228853/2020|EPI_ISL_75341    ····················  ·A··················  ····················  ····················  ····················  ····················
AA    8514 hCoV-19/USA/CA-CHLA-PLM30324031/2020|EPI_ISL_75341    ····················  ·A··················  ····················  ····················  ····················  ····················
AA    8515 hCoV-19/USA/CA-CHLA-PLM58025965/2020|EPI_ISL_75341    ····················  ····················  ····················  ····················  ····················  ····················
AA    8516 hCoV-19/USA/CA-CHLA-PLM14088876/2020|EPI_ISL_75341    ····················  ····················  ····················  ····················  ····················  ····················
AA    8517 hCoV-19/USA/CA-CHLA-PLM26272837/2020|EPI_ISL_75341    ····················  ····················  ····················  ····················  ····················  ····················
AA    8518 hCoV-19/USA/CA-CHLA-PLM88517745/2020|EPI_ISL_75341    ····················  ····················  ····················  ····················  ····················  ····················
AA    8519 hCoV-19/USA/CA-CHLA-PLM74475694/2020|EPI_ISL_75342    ····················  ····················  ····················  ····················  ····················  ····················
AA    8520 hCoV-19/USA/CA-CHLA-PLM40003154/2020|EPI_ISL_75342    ····················  ····················  ····················  ····················  ····················  ····················
AA    8521 hCoV-19/USA/CA-CHLA-PLM36879580/2020|EPI_ISL_75342    ····················  ·A··················  ····················  ····················  ····················  ····················
AA    8522 hCoV-19/USA/CA-CHLA-PLM04548495/2020|EPI_ISL_75342    ····················  ·A··················  ····················  ····················  ····················  ····················
AA    8523 hCoV-19/USA/CA-CHLA-PLM51044374/2020|EPI_ISL_75342    ····················  ·A··················  ····················  ····················  ····················  ····················
AA    8524 hCoV-19/USA/CA-CHLA-PLM04668722/2020|EPI_ISL_75342    ····················  ····················  ····················  ····················  ····················  ····················
AA    8525 hCoV-19/USA/CA-CHLA-PLM31473578/2020|EPI_ISL_75342    ····················  ·A··················  ····················  ····················  ····················  ····················
AA    8526 hCoV-19/USA/CA-CHLA-PLM08517018/2020|EPI_ISL_75342    ····················  ·A··················  ····················  ····················  ····················  ····················
AA    8527 hCoV-19/USA/CA-CHLA-PLM49123428/2020|EPI_ISL_75342    ····················  ·A··················  ····················  ····················  ····················  ····················
AA    8528 hCoV-19/USA/CA-CHLA-PLM51433473/2020|EPI_ISL_75342    ····················  ····················  ····················  ····················  ····················  ····················
AA    8529 hCoV-19/USA/CA-CHLA-PLM06578113/2020|EPI_ISL_75343    ····················  ····················  ····················  ····················  ····················  ····················
AA    8530 hCoV-19/USA/CA-CHLA-PLM82118164/2020|EPI_ISL_75343    ····················  ·A··················  ····················  ····················  ····················  ····················
AA    8531 hCoV-19/USA/CA-CHLA-PLM90211697/2020|EPI_ISL_75343    ····················  ·A··················  ····················  ····················  ····················  ····················
AA    8532 hCoV-19/USA/CA-CHLA-PLM88292776/2020|EPI_ISL_75343    ····················  ····················  ····················  ····················  ····················  ····················
AA    8533 hCoV-19/USA/CA-CHLA-PLM05235743/2020|EPI_ISL_75343    ····················  ····················  ····················  ····················  ····················  ····················
AA    8534 hCoV-19/USA/CA-CHLA-PLM41237673/2020|EPI_ISL_75343    ····················  ····················  ····················  ····················  ····················  ····················
AA    8535 hCoV-19/USA/CA-CHLA-PLM03340521/2020|EPI_ISL_75343    ····················  ·A··················  ····················  ····················  ····················  ····················
AA    8536 hCoV-19/USA/CA-CHLA-PLM40173004/2020|EPI_ISL_75343    ····················  ····················  ····················  ····················  ····················  ····················
AA    8537 hCoV-19/USA/CA-CHLA-PLM64885694/2020|EPI_ISL_75343    ····················  ····················  ····················  ····················  ····················  ····················
AA    8538 hCoV-19/USA/CA-CHLA-PLM52453720/2020|EPI_ISL_75343    ····················  ····················  ····················  ····················  ····················  ····················
AA    8539 hCoV-19/USA/CA-CHLA-PLM50218262/2020|EPI_ISL_75344    ····················  ····················  ····················  ····················  ····················  ····················
AA    8540 hCoV-19/USA/CA-CHLA-PLM47567508/2020|EPI_ISL_75344    ····················  ····················  ····················  ····················  ····················  ····················
AA    8541 hCoV-19/USA/CA-CHLA-PLM41676466/2020|EPI_ISL_75344    ····················  ····················  ····················  ····················  ····················  ····················
AA    8542 hCoV-19/USA/CA-CHLA-PLM87793444/2020|EPI_ISL_75344    ····················  ····················  ····················  ····················  ····················  ····················
AA    8543 hCoV-19/USA/CA-CHLA-PLM12654404/2020|EPI_ISL_75344    ····················  ····················  ····················  ····················  ····················  ····················
AA    8544 hCoV-19/USA/CA-CHLA-PLM23076900/2020|EPI_ISL_75344    ····················  ····················  ····················  ····················  ····················  ····················
AA    8545 hCoV-19/USA/CA-CHLA-PLM07482754/2020|EPI_ISL_75344    ····················  ····················  ····················  ····················  ····················  ····················
AA    8546 hCoV-19/USA/CA-CHLA-PLM57864074/2020|EPI_ISL_75344    ····················  ·A··················  ····················  ····················  ····················  ····················
AA    8547 hCoV-19/USA/CA-CHLA-PLM02905782/2020|EPI_ISL_75344    ····················  ····················  ····················  ····················  ····················  ····················
AA    8548 hCoV-19/USA/CA-CHLA-PLM87677393/2020|EPI_ISL_75344    ····················  ····················  ····················  ····················  ····················  ····················
AA    8549 hCoV-19/USA/CA-CHLA-PLM61959420/2020|EPI_ISL_75345    ····················  ·A··················  ····················  ····················  ····················  ····················
AA    8550 hCoV-19/USA/CA-CHLA-PLM18872077/2020|EPI_ISL_75345    ····················  ·A··················  ····················  ····················  ····················  ····················
AA    8551 hCoV-19/USA/CA-CHLA-PLM90989138/2020|EPI_ISL_75345    ····················  ····················  ····················  ····················  ····················  ····················
AA    8552 hCoV-19/USA/CA-CHLA-PLM57473032/2020|EPI_ISL_75345    ····················  ····················  ····················  ····················  ····················  ····················
AA    8553 hCoV-19/USA/CA-CHLA-PLM88123191/2020|EPI_ISL_75345    ····················  ····················  ····················  ····················  ····················  ····················
AA    8554 hCoV-19/USA/CA-CHLA-PLM74733207/2020|EPI_ISL_75345    ····················  ····················  ····················  ····················  ····················  ····················
AA    8555 hCoV-19/USA/CA-CHLA-PLM29218152/2020|EPI_ISL_75345    ····················  ····················  ····················  ····················  ····················  ····················
AA    8556 hCoV-19/USA/CA-CHLA-PLM21531999/2020|EPI_ISL_75345    ····················  ····················  ····················  ····················  ····················  ····················
AA    8557 hCoV-19/USA/CA-CHLA-PLM58846794/2020|EPI_ISL_75345    ····················  ·A··················  ····················  ····················  ····················  ····················
AA    8558 hCoV-19/USA/CA-CHLA-PLM45816782/2020|EPI_ISL_75345    ····················  ·A··················  ····················  ····················  ····················  ····················
AA    8559 hCoV-19/USA/CA-CHLA-PLM76012104/2020|EPI_ISL_75346    ····················  ····················  ····················  ····················  ····················  ····················
AA    8560 hCoV-19/USA/CA-CHLA-PLM98411854/2020|EPI_ISL_75346    ····················  ····················  ····················  ····················  ····················  ····················
AA    8561 hCoV-19/USA/CA-CHLA-PLM43062990/2020|EPI_ISL_75346    ····················  ····················  ····················  ····················  ····················  ····················
AA    8562 hCoV-19/USA/CA-CHLA-PLM71723340/2020|EPI_ISL_75346    ····················  ····················  ····················  ····················  ····················  ····················
AA    8563 hCoV-19/USA/CA-CHLA-PLM98897037/2020|EPI_ISL_75346    ····················  ·A··················  ····················  ····················  ····················  ····················
AA    8564 hCoV-19/USA/CA-CHLA-PLM33671648/2020|EPI_ISL_75346    ····················  ····················  ····················  ····················  ····················  ····················
AA    8565 hCoV-19/USA/CA-CHLA-PLM69631729/2020|EPI_ISL_75346    ····················  ····················  ····················  ····················  ····················  ····················
AA    8566 hCoV-19/USA/CA-CHLA-PLM45672452/2020|EPI_ISL_75346    ····················  ·A··················  ····················  ····················  ····················  ····················
AA    8567 hCoV-19/USA/CA-CHLA-PLM90547620/2020|EPI_ISL_75346    ····················  ····················  ····················  ····················  ····················  ····················
AA    8568 hCoV-19/USA/CA-CHLA-PLM48900225/2020|EPI_ISL_75346    ····················  ·A··················  ····················  ····················  ····················  ····················
AA    8569 hCoV-19/USA/CA-CHLA-PLM31444580/2020|EPI_ISL_75347    ····················  ····················  ····················  ····················  ····················  ····················
AA    8570 hCoV-19/USA/CA-CHLA-PLM49811241/2020|EPI_ISL_75347    ····················  ·A··················  ····················  ····················  ····················  ····················
AA    8571 hCoV-19/USA/CA-CHLA-PLM61564534/2020|EPI_ISL_75347    ····················  ·A··················  ····················  ····················  ····················  ····················
AA    8572 hCoV-19/USA/CA-CHLA-PLM89293334/2020|EPI_ISL_75347    ····················  ····················  ····················  ····················  ····················  ····················
AA    8573 hCoV-19/USA/CA-CHLA-PLM20572552/2020|EPI_ISL_75347    ····················  ····················  ····················  ····················  ····················  ····················
AA    8574 hCoV-19/USA/CA-CHLA-PLM40051876/2020|EPI_ISL_75347    ····················  ·A··················  ····················  ····················  ····················  ····················
AA    8575 hCoV-19/USA/CA-CHLA-PLM62420958/2020|EPI_ISL_75347    ····················  ····················  ····················  ····················  ····················  ····················
AA    8576 hCoV-19/USA/CA-CHLA-PLM38859034/2020|EPI_ISL_75347    ····················  ·A··················  ····················  ····················  ····················  ····················
AA    8577 hCoV-19/USA/CA-CHLA-PLM58326702/2020|EPI_ISL_75347    ····················  ····················  ····················  ····················  ····················  ····················
AA    8578 hCoV-19/USA/CA-CHLA-PLM02129348/2020|EPI_ISL_75347    ····················  ····················  ····················  ····················  ····················  ····················
AA    8579 hCoV-19/USA/CA-CHLA-PLM84336997/2020|EPI_ISL_75348    ····················  ····················  ····················  ····················  ····················  ····················
AA    8580 hCoV-19/USA/CA-CHLA-PLM43780750/2020|EPI_ISL_75348    ····················  ····················  ····················  ····················  ····················  ····················
AA    8581 hCoV-19/USA/CA-CHLA-PLM53657240/2020|EPI_ISL_75348    ····················  ····················  ····················  ····················  ····················  ····················
AA    8582 hCoV-19/USA/CA-CHLA-PLM76781771/2020|EPI_ISL_75348    ····················  ·A··················  ····················  ····················  ····················  ····················
AA    8583 hCoV-19/USA/CA-CHLA-PLM03763789/2020|EPI_ISL_75348    ····················  ····················  ····················  ····················  ····················  ····················
AA    8584 hCoV-19/USA/CA-CHLA-PLM42821873/2020|EPI_ISL_75348    ····················  ····················  ····················  ····················  ····················  ····················
AA    8585 hCoV-19/USA/CA-CHLA-PLM81568971/2020|EPI_ISL_75348    ····················  ·A··················  ····················  ····················  ····················  ····················
AA    8586 hCoV-19/USA/CA-CHLA-PLM65528193/2020|EPI_ISL_75348    ····················  ····················  ····················  ····················  ····················  ····················
AA    8587 hCoV-19/USA/CA-CHLA-PLM86862197/2020|EPI_ISL_75348    ····················  ·A··················  ····················  ····················  ····················  ····················
AA    8588 hCoV-19/USA/CA-CHLA-PLM00143709/2020|EPI_ISL_75349    ····················  ·A··················  ····················  ····················  ····················  ····················
AA    8589 hCoV-19/USA/CA-CHLA-PLM52743522/2020|EPI_ISL_75349    ····················  ····················  ····················  ····················  ····················  ····················
```

| | | | | | | | |
|---|---|---|---|---|---|---|---|
| AA | 8590 hCoV-19/USA/CA-CHLA-PLM65511564/2020\|EPI_ISL_75349 | ···················· | ···················· | ···················· | ···················· | ···················· | ···················· |
| AA | 8591 hCoV-19/USA/CA-CHLA-PLM92698286/2020\|EPI_ISL_75349 | ···················· | ·A·················· | ···················· | ···················· | ···················· | ···················· |
| AA | 8592 hCoV-19/USA/CA-CHLA-PLM81753386/2020\|EPI_ISL_75349 | ···················· | ·A·················· | ···················· | ···················· | ···················· | ···················· |
| AA | 8593 hCoV-19/USA/CA-CHLA-PLM24615260/2020\|EPI_ISL_75349 | ···················· | ···················· | ···················· | ···················· | ···················· | ···················· |
| AA | 8594 hCoV-19/USA/CA-CHLA-PLM16398520/2020\|EPI_ISL_75349 | ···················· | ···················· | ···················· | ···················· | ···················· | ···················· |
| AA | 8595 hCoV-19/USA/NY-Wadsworth-248114-01/2020\|EPI_ISL_75 | ···················· | ···················· | ···················· | ···················· | ···················· | ···················· |
| AA | 8596 hCoV-19/USA/NY-Wadsworth-248111-01/2020\|EPI_ISL_75 | ···················· | ···················· | ···················· | ···················· | ···················· | ···················· |
| AA | 8597 hCoV-19/USA/NY-Wadsworth-248245-01/2020\|EPI_ISL_75 | ···················· | ···················· | ···················· | ···················· | ···················· | ···················· |
| AA | 8598 hCoV-19/USA/NY-Wadsworth-247821-01/2020\|EPI_ISL_75 | ···················· | ···················· | ···················· | ···················· | ···················· | ···················· |
| AA | 8599 hCoV-19/USA/NY-Wadsworth-247823-01/2020\|EPI_ISL_75 | ···················· | ···················· | ···················· | ···················· | ···················· | ···················· |
| AA | 8600 hCoV-19/USA/NY-Wadsworth-247827-01/2020\|EPI_ISL_75 | ···················· | ···················· | ···················· | ···················· | ···················· | ···················· |
| AA | 8601 hCoV-19/USA/NY-Wadsworth-248039-01/2020\|EPI_ISL_75 | ···················· | ···················· | ···················· | ···················· | ···················· | ···················· |
| AA | 8602 hCoV-19/USA/NY-Wadsworth-248282-01/2020\|EPI_ISL_75 | ···················· | ···················· | ···················· | ···················· | ···················· | ···················· |
| AA | 8603 hCoV-19/USA/NY-Wadsworth-247810-01/2020\|EPI_ISL_75 | ···················· | ···················· | ···················· | ···················· | ···················· | ···················· |
| AA | 8604 hCoV-19/USA/NY-Wadsworth-247813-01/2020\|EPI_ISL_75 | ···················· | ···················· | ···················· | ···················· | ···················· | ···················· |
| AA | 8605 hCoV-19/USA/NY-Wadsworth-248019-01/2020\|EPI_ISL_75 | ···················· | ···················· | ···················· | ···················· | ···················· | ···················· |
| AA | 8606 hCoV-19/USA/NY-Wadsworth-248026-01/2020\|EPI_ISL_75 | ···················· | ···················· | ···················· | ···················· | ···················· | ···················· |
| AA | 8607 hCoV-19/USA/NY-Wadsworth-248592-01/2020\|EPI_ISL_75 | ···················· | ···················· | ···················· | ···················· | ···················· | ···················· |
| AA | 8608 hCoV-19/USA/NY-Wadsworth-248683-01/2020\|EPI_ISL_75 | ···················· | ···················· | ···················· | ···················· | ···················· | ···················· |
| AA | 8609 hCoV-19/USA/NY-Wadsworth-248623-01/2020\|EPI_ISL_75 | ···················· | ···················· | ···················· | ···················· | ···················· | ···················· |
| AA | 8610 hCoV-19/USA/NY-Wadsworth-248740-01/2020\|EPI_ISL_75 | ···················· | ···················· | ···················· | ···················· | ···················· | ···················· |
| AA | 8611 hCoV-19/USA/NY-Wadsworth-248637-01/2020\|EPI_ISL_75 | ···················· | ···················· | ···················· | ···················· | ···················· | ···················· |
| AA | 8612 hCoV-19/USA/NY-Wadsworth-248646-01/2020\|EPI_ISL_75 | ···················· | ···················· | ···················· | ···················· | ···················· | ···················· |
| AA | 8613 hCoV-19/USA/NY-Wadsworth-248654-01/2020\|EPI_ISL_75 | ···················· | ···················· | ···················· | ···················· | ···················· | ···················· |
| AA | 8614 hCoV-19/USA/NY-Wadsworth-248655-01/2020\|EPI_ISL_75 | ···················· | ···················· | ···················· | ···················· | ···················· | ···················· |
| AA | 8615 hCoV-19/USA/NY-Wadsworth-248657-01/2020\|EPI_ISL_75 | ···················· | ···················· | ···················· | ···················· | ···················· | ···················· |
| AA | 8616 hCoV-19/USA/NY-Wadsworth-248584-01/2020\|EPI_ISL_75 | ···················· | ···················· | ···················· | ···················· | ···················· | ···················· |
| AA | 8617 hCoV-19/USA/NY-Wadsworth-248784-01/2020\|EPI_ISL_75 | ···················· | ···················· | ···················· | ···················· | ···················· | ···················· |
| AA | 8618 hCoV-19/USA/NY-Wadsworth-248767-01/2020\|EPI_ISL_75 | ···················· | ···················· | ···················· | ···················· | ···················· | ···················· |
| AA | 8619 hCoV-19/USA/NY-Wadsworth-249516-01/2020\|EPI_ISL_75 | ···················· | ·A·················· | ···················· | ···················· | ···················· | ···················· |
| AA | 8620 hCoV-19/USA/NY-Wadsworth-249555-01/2020\|EPI_ISL_75 | ···················· | ·A·················· | ···················· | ···················· | ···················· | ···················· |
| AA | 8621 hCoV-19/USA/NY-Wadsworth-249744-01/2020\|EPI_ISL_75 | ···················· | ···················· | ···················· | ···················· | ···················· | ···················· |
| AA | 8622 hCoV-19/USA/NY-Wadsworth-249749-01/2020\|EPI_ISL_75 | ···················· | ···················· | ···················· | ···················· | ···················· | ···················· |
| AA | 8623 hCoV-19/USA/NY-Wadsworth-249750-01/2020\|EPI_ISL_75 | ···················· | ···················· | ···················· | ···················· | ···················· | ···················· |
| AA | 8624 hCoV-19/USA/NY-Wadsworth-249754-01/2020\|EPI_ISL_75 | ···················· | ···················· | ···················· | ···················· | ···················· | ···················· |
| AA | 8625 hCoV-19/USA/NY-Wadsworth-249828-01/2020\|EPI_ISL_75 | ···················· | ···················· | ···················· | ···················· | ···················· | ···················· |
| AA | 8626 hCoV-19/USA/NY-Wadsworth-249945-01/2020\|EPI_ISL_75 | ···················· | ···················· | ···················· | ···················· | ···················· | ···················· |
| AA | 8627 hCoV-19/USA/NY-Wadsworth-250294-01/2020\|EPI_ISL_75 | ···················· | ···················· | ···················· | ···················· | ···················· | ···················· |
| AA | 8628 hCoV-19/USA/NY-Wadsworth-249769-01/2020\|EPI_ISL_75 | ···················· | ···················· | ···················· | ···················· | ···················· | ···················· |
| AA | 8629 hCoV-19/USA/NY-Wadsworth-249770-01/2020\|EPI_ISL_75 | ···················· | ···················· | ···················· | ···················· | ···················· | ···················· |
| AA | 8630 hCoV-19/USA/NY-Wadsworth-249773-01/2020\|EPI_ISL_75 | ···················· | ···················· | ···················· | ···················· | ···················· | ···················· |
| AA | 8631 hCoV-19/USA/NY-Wadsworth-249779-01/2020\|EPI_ISL_75 | ···················· | ···················· | ···················· | ···················· | ···················· | ···················· |
| AA | 8632 hCoV-19/USA/NY-Wadsworth-249783-01/2020\|EPI_ISL_75 | ···················· | ···················· | ···················· | ···················· | ···················· | ···················· |
| AA | 8633 hCoV-19/USA/NY-Wadsworth-249787-01/2020\|EPI_ISL_75 | ···················· | ···················· | ···················· | ···················· | ···················· | ···················· |
| AA | 8634 hCoV-19/USA/NY-Wadsworth-249890-01/2020\|EPI_ISL_75 | ···················· | ···················· | ···················· | ···················· | ···················· | ···················· |
| AA | 8635 hCoV-19/USA/NY-Wadsworth-249912-01/2020\|EPI_ISL_75 | ···················· | ···················· | ···················· | ···················· | ···················· | ···················· |
| AA | 8636 hCoV-19/USA/NY-Wadsworth-249933-01/2020\|EPI_ISL_75 | ···················· | ···················· | ···················· | ···················· | ···················· | ···················· |
| AA | 8637 hCoV-19/USA/NY-Wadsworth-249934-01/2020\|EPI_ISL_75 | ···················· | ···················· | ···················· | ···················· | ···················· | ···················· |
| AA | 8638 hCoV-19/USA/NY-Wadsworth-251941-01/2020\|EPI_ISL_75 | ···················· | ·A·················· | ···················· | ···················· | ···················· | ···················· |
| AA | 8639 hCoV-19/USA/NY-Wadsworth-251938-01/2020\|EPI_ISL_75 | ···················· | ·A·················· | ···················· | ···················· | ···················· | ···················· |
| AA | 8640 hCoV-19/USA/GA-EHC-033G/2020\|EPI_ISL_754751\|2020-0 | ···················· | ···················· | ···················· | ···················· | ···················· | ···················· |
| AA | 8641 hCoV-19/USA/GA-EHC-034H/2020\|EPI_ISL_754752\|2020-0 | ···················· | ···················· | ···················· | ···················· | ···················· | ···················· |
| AA | 8642 hCoV-19/USA/GA-EHC-036J/2020\|EPI_ISL_754753\|2020-0 | ···················· | ···················· | ···················· | ···················· | ···················· | ···················· |
| AA | 8643 hCoV-19/USA/GA-EHC-037K/2020\|EPI_ISL_754754\|2020-0 | ···················· | ···················· | ···················· | ···················· | ···················· | ···················· |
| AA | 8644 hCoV-19/USA/GA-EHC-038L/2020\|EPI_ISL_754755\|2020-0 | ···················· | ···················· | ···················· | ···················· | ···················· | ···················· |
| AA | 8645 hCoV-19/USA/GA-EHC-040N/2020\|EPI_ISL_754756\|2020-0 | ···················· | ···················· | ···················· | ···················· | ···················· | ···················· |
| AA | 8646 hCoV-19/USA/GA-EHC-041O/2020\|EPI_ISL_754757\|2020-0 | ···················· | ···················· | ···················· | ···················· | ···················· | ···················· |
| AA | 8647 hCoV-19/USA/GA-EHC-042P/2020\|EPI_ISL_754758\|2020-0 | ···················· | ···················· | ···················· | ···················· | ···················· | ···················· |
| AA | 8648 hCoV-19/USA/GA-EHC-043Q/2020\|EPI_ISL_754759\|2020-0 | ···················· | ···················· | ···················· | ···················· | ···················· | ···················· |
| AA | 8649 hCoV-19/USA/GA-EHC-044R/2020\|EPI_ISL_754760\|2020-0 | ···················· | ···················· | ···················· | ···················· | ···················· | ···················· |
| AA | 8650 hCoV-19/USA/GA-EHC-049W/2020\|EPI_ISL_754761\|2020-0 | ···················· | ···················· | ···················· | ···················· | ···················· | ···················· |
| AA | 8651 hCoV-19/USA/GA-EHC-052Z/2020\|EPI_ISL_754762\|2020-0 | ···················· | ···················· | ···················· | ···················· | ···················· | ···················· |
| AA | 8652 hCoV-19/USA/GA-EHC-053A/2020\|EPI_ISL_754763\|2020-0 | ···················· | ···················· | ···················· | ···················· | ···················· | ···················· |
| AA | 8653 hCoV-19/USA/GA-EHC-054B/2020\|EPI_ISL_754764\|2020-0 | ···················· | ···················· | ···················· | ···················· | ···················· | ···················· |
| AA | 8654 hCoV-19/USA/GA-EHC-055C/2020\|EPI_ISL_754765\|2020-0 | ···················· | ···················· | ···················· | ···················· | ···················· | ···················· |
| AA | 8655 hCoV-19/USA/GA-EHC-097S/2020\|EPI_ISL_754766\|2020-0 | ···················· | ···················· | ···················· | ···················· | ···················· | ···················· |
| AA | 8656 hCoV-19/USA/GA-EHC-110F/2020\|EPI_ISL_754767\|2020-0 | ···················· | ···················· | ···················· | ···················· | ···················· | ···················· |
| AA | 8657 hCoV-19/USA/GA-EHC-113J/2020\|EPI_ISL_754768\|2020-0 | ···················· | ···················· | ···················· | ···················· | ···················· | ···················· |
| AA | 8658 hCoV-19/USA/GA-EHC-139K/2020\|EPI_ISL_754769\|2020-0 | ···················· | ···················· | ···················· | ···················· | ···················· | ···················· |
| AA | 8659 hCoV-19/USA/GA-EHC-419E/2020\|EPI_ISL_754770\|2020-1 | ···················· | ···A················ | ···················· | ···················· | ···················· | ···················· |
| AA | 8660 hCoV-19/USA/GA-EHC-415A/2020\|EPI_ISL_754771\|2020-1 | ···················· | ···················· | ···················· | ···················· | ···················· | ···················· |
| AA | 8661 hCoV-19/USA/GA-EHC-417C/2020\|EPI_ISL_754772\|2020-1 | ···················· | ···················· | ···················· | ···················· | ···················· | ···················· |
| AA | 8662 hCoV-19/USA/GA-EHC-475I/2020\|EPI_ISL_754773\|2020-1 | ···················· | ···················· | ···················· | ···················· | ···················· | ···················· |
| AA | 8663 hCoV-19/USA/GA-EHC-476J/2020\|EPI_ISL_754774\|2020-1 | ···················· | ···················· | ···················· | ···················· | ···················· | ···················· |
| AA | 8664 hCoV-19/USA/GA-EHC-472F/2020\|EPI_ISL_754775\|2020-1 | ···················· | ···················· | ···················· | ···················· | ···················· | ···················· |
| AA | 8665 hCoV-19/USA/GA-EHC-023W/2020\|EPI_ISL_754776\|2020-0 | ···················· | ···················· | ···················· | ···················· | ···················· | ···················· |
| AA | 8666 hCoV-19/USA/GA-EHC-060H/2020\|EPI_ISL_754777\|2020-0 | ···················· | ···················· | ···················· | ···················· | ···················· | ···················· |
| AA | 8667 hCoV-19/USA/GA-EHC-064L/2020\|EPI_ISL_754778\|2020-0 | ···················· | ···················· | ···················· | ···················· | ···················· | ···················· |
| AA | 8668 hCoV-19/Denmark/DCGC-21586/2020\|EPI_ISL_758250\|202 | ···················· | ···················· | ···················· | ···················· | ···················· | ···················· |
| AA | 8669 hCoV-19/Denmark/DCGC-21588/2020\|EPI_ISL_758251\|202 | ···················· | ···················· | ···················· | ···················· | ···················· | ···················· |

| | | | | | | | |
|---|---|---|---|---|---|---|---|
| AA | 8670 hCoV-19/Denmark/DCGC-21589/2020\|EPI_ISL_758252\|202 | .................... | .................... | .................... | .................... | .................... | .................... |
| AA | 8671 hCoV-19/Denmark/DCGC-21592/2020\|EPI_ISL_758253\|202 | .................... | .................... | .................... | .................... | .................... | .................... |
| AA | 8672 hCoV-19/Denmark/DCGC-21593/2020\|EPI_ISL_758254\|202 | .................... | .................... | .................... | .................... | .................... | .................... |
| AA | 8673 hCoV-19/Denmark/DCGC-21597/2020\|EPI_ISL_758255\|202 | .................... | .................... | .................... | .................... | .................... | .................... |
| AA | 8674 hCoV-19/Denmark/DCGC-21600/2020\|EPI_ISL_758256\|202 | .................... | .................... | .................... | .................... | .................... | .................... |
| AA | 8675 hCoV-19/Denmark/DCGC-21602/2020\|EPI_ISL_758258\|202 | .................... | .................... | .................... | .................... | .................... | .................... |
| AA | 8676 hCoV-19/Denmark/DCGC-21607/2020\|EPI_ISL_758260\|202 | .................... | .................... | .................... | .................... | .................... | .................... |
| AA | 8677 hCoV-19/Denmark/DCGC-21609/2020\|EPI_ISL_758261\|202 | .................... | .................... | .................... | .................... | .................... | .................... |
| AA | 8678 hCoV-19/Denmark/DCGC-21610/2020\|EPI_ISL_758262\|202 | .................... | .................... | .................... | .................... | .................... | .................... |
| AA | 8679 hCoV-19/Denmark/DCGC-21613/2020\|EPI_ISL_758263\|202 | .................... | .................... | .................... | .................... | .................... | .................... |
| AA | 8680 hCoV-19/Denmark/DCGC-21614/2020\|EPI_ISL_758264\|202 | .................... | .................... | .................... | .................... | .................... | .................... |
| AA | 8681 hCoV-19/Denmark/DCGC-21618/2020\|EPI_ISL_758265\|202 | .................... | .................... | .................... | .................... | .................... | .................... |
| AA | 8682 hCoV-19/Denmark/DCGC-21619/2020\|EPI_ISL_758266\|202 | .................... | .................... | .................... | .................... | .................... | .................... |
| AA | 8683 hCoV-19/Denmark/DCGC-21620/2020\|EPI_ISL_758267\|202 | .................... | .................... | .................... | .................... | .................... | .................... |
| AA | 8684 hCoV-19/Denmark/DCGC-21623/2020\|EPI_ISL_758268\|202 | .................... | .................... | .................... | .................... | .................... | .................... |
| AA | 8685 hCoV-19/Denmark/DCGC-21624/2020\|EPI_ISL_758269\|202 | .................... | .................... | .................... | .................... | .................... | .................... |
| AA | 8686 hCoV-19/Denmark/DCGC-21155/2020\|EPI_ISL_759580\|202 | .................... | .................... | .................... | .................... | .................... | .................... |
| AA | 8687 hCoV-19/Denmark/DCGC-21158/2020\|EPI_ISL_759581\|202 | .................... | .................... | .................... | .................... | .................... | .................... |
| AA | 8688 hCoV-19/Denmark/DCGC-21159/2020\|EPI_ISL_759582\|202 | .................... | .................... | .................... | .................... | .................... | .................... |
| AA | 8689 hCoV-19/Denmark/DCGC-21160/2020\|EPI_ISL_759583\|202 | .................... | .................... | .................... | .................... | .................... | .................... |
| AA | 8690 hCoV-19/Denmark/DCGC-21161/2020\|EPI_ISL_759584\|202 | .................... | .................... | .................... | .................... | .................... | .................... |
| AA | 8691 hCoV-19/Denmark/DCGC-21162/2020\|EPI_ISL_759585\|202 | .................... | .................... | .................... | .................... | .................... | .................... |
| AA | 8692 hCoV-19/Denmark/DCGC-21164/2020\|EPI_ISL_759586\|202 | .................... | .................... | .................... | .................... | .................... | .................... |
| AA | 8693 hCoV-19/Denmark/DCGC-21167/2020\|EPI_ISL_759587\|202 | .................... | .................... | .................... | .................... | .................... | .................... |
| AA | 8694 hCoV-19/Denmark/DCGC-21170/2020\|EPI_ISL_759589\|202 | .................... | .................... | .................... | .................... | .................... | .................... |
| AA | 8695 hCoV-19/Denmark/DCGC-21172/2020\|EPI_ISL_759590\|202 | .................... | .................... | .................... | .................... | .................... | .................... |
| AA | 8696 hCoV-19/Denmark/DCGC-21173/2020\|EPI_ISL_759591\|202 | .................... | .................... | .................... | .................... | .................... | .................... |
| AA | 8697 hCoV-19/Denmark/DCGC-21174/2020\|EPI_ISL_759592\|202 | .................... | .................... | .................... | .................... | .................... | .................... |
| AA | 8698 hCoV-19/Denmark/DCGC-21175/2020\|EPI_ISL_759593\|202 | .................... | .................... | .................... | .................... | .................... | .................... |
| AA | 8699 hCoV-19/Denmark/DCGC-21177/2020\|EPI_ISL_759594\|202 | .................... | .................... | .................... | .................... | .................... | .................... |
| AA | 8700 hCoV-19/Denmark/DCGC-21178/2020\|EPI_ISL_759595\|202 | .................... | .................... | .................... | .................... | .................... | .................... |
| AA | 8701 hCoV-19/Denmark/DCGC-21180/2020\|EPI_ISL_759596\|202 | .................... | .................... | .................... | .................... | .................... | .................... |
| AA | 8702 hCoV-19/Denmark/DCGC-21181/2020\|EPI_ISL_759597\|202 | .................... | .................... | .................... | .................... | .................... | .................... |
| AA | 8703 hCoV-19/Denmark/DCGC-21182/2020\|EPI_ISL_759598\|202 | .................... | .................... | .................... | .................... | .................... | .................... |
| AA | 8704 hCoV-19/Denmark/DCGC-21183/2020\|EPI_ISL_759599\|202 | .................... | .................... | .................... | .................... | .................... | .................... |
| AA | 8705 hCoV-19/Denmark/DCGC-19942/2020\|EPI_ISL_747617\|202 | .................... | .................... | .................... | .................... | .................... | .................... |
| AA | 8706 hCoV-19/Denmark/DCGC-19992/2020\|EPI_ISL_747618\|202 | .................... | .................... | .................... | .................... | .................... | .................... |
| AA | 8707 hCoV-19/Denmark/DCGC-20100/2020\|EPI_ISL_747619\|202 | .................... | .................... | .................... | .................... | .................... | .................... |
| AA | 8708 hCoV-19/Denmark/DCGC-20121/2020\|EPI_ISL_747620\|202 | .................... | .................... | .................... | .................... | .................... | .................... |
| AA | 8709 hCoV-19/Denmark/DCGC-20134/2020\|EPI_ISL_747621\|202 | .................... | .................... | .................... | .................... | .................... | .................... |
| AA | 8710 hCoV-19/Denmark/DCGC-20154/2020\|EPI_ISL_747622\|202 | .................... | .................... | .................... | .................... | .................... | .................... |
| AA | 8711 hCoV-19/Denmark/DCGC-20160/2020\|EPI_ISL_747623\|202 | .................... | .................... | .................... | .................... | .................... | .................... |
| AA | 8712 hCoV-19/Denmark/DCGC-20206/2020\|EPI_ISL_747624\|202 | .................... | .................... | .................... | .................... | .................... | .................... |
| AA | 8713 hCoV-19/Denmark/DCGC-20227/2020\|EPI_ISL_747625\|202 | .................... | .................... | .................... | .................... | .................... | .................... |
| AA | 8714 hCoV-19/Denmark/DCGC-20256/2020\|EPI_ISL_747626\|202 | .................... | .................... | .................... | .................... | .................... | .................... |
| AA | 8715 hCoV-19/Denmark/DCGC-20260/2020\|EPI_ISL_747627\|202 | .................... | .................... | .................... | .................... | .................... | .................... |
| AA | 8716 hCoV-19/Denmark/DCGC-20288/2020\|EPI_ISL_747628\|202 | .................... | .................... | .................... | .................... | .................... | .................... |
| AA | 8717 hCoV-19/Denmark/DCGC-20301/2020\|EPI_ISL_747629\|202 | .................... | .................... | .................... | .................... | .................... | .................... |
| AA | 8718 hCoV-19/Denmark/DCGC-20309/2020\|EPI_ISL_747630\|202 | .................... | .................... | .................... | .................... | .................... | .................... |
| AA | 8719 hCoV-19/Denmark/DCGC-20311/2020\|EPI_ISL_747631\|202 | .................... | .................... | .................... | .................... | .................... | .................... |
| AA | 8720 hCoV-19/Denmark/DCGC-19902/2020\|EPI_ISL_747632\|202 | .................... | .................... | .................... | .................... | .................... | .................... |
| AA | 8721 hCoV-19/Denmark/DCGC-20165/2020\|EPI_ISL_747633\|202 | .................... | .................... | .................... | .................... | .................... | .................... |
| AA | 8722 hCoV-19/Denmark/DCGC-20223/2020\|EPI_ISL_747634\|202 | .................... | .................... | .................... | .................... | .................... | .................... |
| AA | 8723 hCoV-19/Denmark/DCGC-20272/2020\|EPI_ISL_747635\|202 | .................... | .................... | .................... | .................... | .................... | .................... |
| AA | 8724 hCoV-19/Denmark/DCGC-19765/2020\|EPI_ISL_747661\|202 | .................... | .................... | .................... | .................... | .................... | .................... |
| AA | 8725 hCoV-19/Denmark/DCGC-19795/2020\|EPI_ISL_747662\|202 | .................... | .................... | .................... | .................... | .................... | .................... |
| AA | 8726 hCoV-19/Denmark/DCGC-19836/2020\|EPI_ISL_747663\|202 | .................... | .................... | .................... | .................... | .................... | .................... |
| AA | 8727 hCoV-19/Denmark/DCGC-19847/2020\|EPI_ISL_747664\|202 | .................... | .................... | .................... | .................... | .................... | .................... |
| AA | 8728 hCoV-19/Denmark/DCGC-19935/2020\|EPI_ISL_747665\|202 | .................... | .................... | .................... | .................... | .................... | .................... |
| AA | 8729 hCoV-19/Denmark/DCGC-19953/2020\|EPI_ISL_747666\|202 | .................... | .................... | .................... | .................... | .................... | .................... |
| AA | 8730 hCoV-19/Denmark/DCGC-20057/2020\|EPI_ISL_747667\|202 | .................... | .................... | .................... | .................... | .................... | .................... |
| AA | 8731 hCoV-19/Denmark/DCGC-20071/2020\|EPI_ISL_747668\|202 | .................... | .................... | .................... | .................... | .................... | .................... |
| AA | 8732 hCoV-19/Denmark/DCGC-20080/2020\|EPI_ISL_747669\|202 | .................... | .................... | .................... | .................... | .................... | .................... |
| AA | 8733 hCoV-19/Denmark/DCGC-20085/2020\|EPI_ISL_747670\|202 | .................... | .................... | .................... | .................... | .................... | .................... |
| AA | 8734 hCoV-19/Denmark/DCGC-20109/2020\|EPI_ISL_747671\|202 | .................... | .................... | .................... | .................... | .................... | .................... |
| AA | 8735 hCoV-19/Denmark/DCGC-20118/2020\|EPI_ISL_747672\|202 | .................... | .................... | .................... | .................... | .................... | .................... |
| AA | 8736 hCoV-19/Denmark/DCGC-20130/2020\|EPI_ISL_747673\|202 | .................... | .................... | .................... | .................... | .................... | .................... |
| AA | 8737 hCoV-19/Denmark/DCGC-20153/2020\|EPI_ISL_747674\|202 | .................... | .................... | .................... | .................... | .................... | .................... |
| AA | 8738 hCoV-19/Denmark/DCGC-20271/2020\|EPI_ISL_747675\|202 | .................... | .................... | .................... | .................... | .................... | .................... |
| AA | 8739 hCoV-19/Denmark/DCGC-20277/2020\|EPI_ISL_747676\|202 | .................... | .................... | .................... | .................... | .................... | .................... |
| AA | 8740 hCoV-19/Denmark/DCGC-20280/2020\|EPI_ISL_747677\|202 | .................... | .................... | .................... | .................... | .................... | .................... |
| AA | 8741 hCoV-19/Denmark/DCGC-20281/2020\|EPI_ISL_747678\|202 | .................... | .................... | .................... | .................... | .................... | .................... |
| AA | 8742 hCoV-19/Denmark/DCGC-20292/2020\|EPI_ISL_747679\|202 | .................... | .................... | .................... | .................... | .................... | .................... |
| AA | 8743 hCoV-19/Denmark/DCGC-20299/2020\|EPI_ISL_747680\|202 | .................... | .................... | .................... | .................... | .................... | .................... |
| AA | 8744 hCoV-19/Denmark/DCGC-19794/2020\|EPI_ISL_747681\|202 | .................... | .................... | .................... | .................... | .................... | .................... |
| AA | 8745 hCoV-19/Denmark/DCGC-19802/2020\|EPI_ISL_747682\|202 | .................... | .................... | .................... | .................... | .................... | .................... |
| AA | 8746 hCoV-19/Denmark/DCGC-19808/2020\|EPI_ISL_747683\|202 | .................... | .................... | .................... | .................... | .................... | .................... |
| AA | 8747 hCoV-19/Denmark/DCGC-19815/2020\|EPI_ISL_747684\|202 | .................... | .................... | .................... | .................... | .................... | .................... |
| AA | 8748 hCoV-19/Denmark/DCGC-19824/2020\|EPI_ISL_747685\|202 | .................... | .................... | .................... | .................... | .................... | .................... |
| AA | 8749 hCoV-19/Denmark/DCGC-19849/2020\|EPI_ISL_747686\|202 | .................... | .................... | .................... | .................... | .................... | .................... |

AA 8750 hCoV-19/Denmark/DCGC-19865/2020|EPI_ISL_747687|202 .................... .................... .................... .................... .................... ....................
AA 8751 hCoV-19/Denmark/DCGC-19880/2020|EPI_ISL_747688|202 .................... .................... .................... .................... .................... ....................
AA 8752 hCoV-19/Denmark/DCGC-19884/2020|EPI_ISL_747689|202 .................... .................... .................... .................... .................... ....................
AA 8753 hCoV-19/Denmark/DCGC-19888/2020|EPI_ISL_747690|202 .................... .................... .................... .................... .................... ....................
AA 8754 hCoV-19/Denmark/DCGC-19892/2020|EPI_ISL_747691|202 .................... .................... .................... .................... .................... ....................
AA 8755 hCoV-19/Denmark/DCGC-19895/2020|EPI_ISL_747692|202 .................... .................... .................... .................... .................... ....................
AA 8756 hCoV-19/Denmark/DCGC-19898/2020|EPI_ISL_747693|202 .................... .................... .................... .................... .................... ....................
AA 8757 hCoV-19/Denmark/DCGC-19918/2020|EPI_ISL_747694|202 .................... .................... .................... .................... .................... ....................
AA 8758 hCoV-19/Denmark/DCGC-19954/2020|EPI_ISL_747695|202 .................... .................... .................... .................... .................... ....................
AA 8759 hCoV-19/Denmark/DCGC-19973/2020|EPI_ISL_747696|202 .................... .................... .................... .................... .................... ....................
AA 8760 hCoV-19/Denmark/DCGC-20002/2020|EPI_ISL_747697|202 .................... .................... .................... .................... .................... ....................
AA 8761 hCoV-19/Denmark/DCGC-20012/2020|EPI_ISL_747698|202 .................... .................... .................... .................... .................... ....................
AA 8762 hCoV-19/Denmark/DCGC-20030/2020|EPI_ISL_747699|202 .................... .................... .................... .................... .................... ....................
AA 8763 hCoV-19/Denmark/DCGC-19364/2020|EPI_ISL_748906|202 .................... .................... .................... .................... .................... ....................
AA 8764 hCoV-19/Denmark/DCGC-19365/2020|EPI_ISL_748907|202 .................... .................... .................... .................... .................... ....................
AA 8765 hCoV-19/Denmark/DCGC-19440/2020|EPI_ISL_748948|202 .................... .................... .................... .................... .................... ....................
AA 8766 hCoV-19/Denmark/DCGC-19441/2020|EPI_ISL_748949|202 .................... .................... .................... .................... .................... ....................
AA 8767 hCoV-19/Denmark/DCGC-19442/2020|EPI_ISL_748950|202 .................... .................... .................... .................... .................... ....................
AA 8768 hCoV-19/Denmark/DCGC-19443/2020|EPI_ISL_748951|202 .................... .................... .................... .................... .................... ....................
AA 8769 hCoV-19/Denmark/DCGC-19447/2020|EPI_ISL_748952|202 .................... .................... .................... .................... .................... ....................
AA 8770 hCoV-19/Denmark/DCGC-19448/2020|EPI_ISL_748953|202 .................... .................... .................... .................... .................... ....................
AA 8771 hCoV-19/Denmark/DCGC-19449/2020|EPI_ISL_748954|202 .................... .................... .................... .................... .................... ....................
AA 8772 hCoV-19/Denmark/DCGC-19450/2020|EPI_ISL_748955|202 .................... .................... .................... .................... .................... ....................
AA 8773 hCoV-19/Denmark/DCGC-19452/2020|EPI_ISL_748956|202 .................... .................... .................... .................... .................... ....................
AA 8774 hCoV-19/Denmark/DCGC-19454/2020|EPI_ISL_748957|202 .................... .................... .................... .................... .................... ....................
AA 8775 hCoV-19/Denmark/DCGC-19455/2020|EPI_ISL_748958|202 .................... .A.................. .................... .................... .................... ...A................
AA 8776 hCoV-19/Denmark/DCGC-19456/2020|EPI_ISL_748959|202 .................... .................... .................... .................... .................... ....................
AA 8777 hCoV-19/Denmark/DCGC-19457/2020|EPI_ISL_748960|202 .................... .................... .................... .................... .................... ....................
AA 8778 hCoV-19/Denmark/DCGC-19460/2020|EPI_ISL_748961|202 .................... .................... .................... .................... .................... ....................
AA 8779 hCoV-19/Denmark/DCGC-19462/2020|EPI_ISL_748962|202 .................... .................... .................... .................... .................... ....................
AA 8780 hCoV-19/Denmark/DCGC-19464/2020|EPI_ISL_748963|202 .................... .................... .................... .................... .................... ....................
AA 8781 hCoV-19/Denmark/DCGC-19465/2020|EPI_ISL_748964|202 .................... .................... .................... .................... .................... ....................
AA 8782 hCoV-19/Denmark/DCGC-19466/2020|EPI_ISL_748965|202 .................... .................... .................... .................... .................... ....................
AA 8783 hCoV-19/Denmark/DCGC-19467/2020|EPI_ISL_748966|202 .................... .................... .................... .................... .................... ....................
AA 8784 hCoV-19/Denmark/DCGC-19468/2020|EPI_ISL_748967|202 .................... .................... .................... .................... .................... ....................
AA 8785 hCoV-19/Denmark/DCGC-19469/2020|EPI_ISL_748968|202 .................... .................... .................... .................... .................... ....................
AA 8786 hCoV-19/Denmark/DCGC-19470/2020|EPI_ISL_748969|202 .................... .................... .................... .................... .................... ....................
AA 8787 hCoV-19/Denmark/DCGC-19471/2020|EPI_ISL_748970|202 .................... .................... .................... .................... .................... ....................
AA 8788 hCoV-19/Denmark/DCGC-19472/2020|EPI_ISL_748971|202 .................... .A.................. .................... .................... .................... ....................
AA 8789 hCoV-19/Denmark/DCGC-19473/2020|EPI_ISL_748972|202 .................... .................... .................... .................... .................... ....................
AA 8790 hCoV-19/Denmark/DCGC-19474/2020|EPI_ISL_748973|202 .................... .................... .................... .................... .................... ....................
AA 8791 hCoV-19/Denmark/DCGC-19475/2020|EPI_ISL_748974|202 .................... .................... .................... .................... .................... ....................
AA 8792 hCoV-19/Denmark/DCGC-19476/2020|EPI_ISL_748975|202 .................... .A.................. .................... .................... .................... ....................
AA 8793 hCoV-19/Denmark/DCGC-19477/2020|EPI_ISL_748976|202 .................... .................... .................... .................... .................... ....................
AA 8794 hCoV-19/Denmark/DCGC-19478/2020|EPI_ISL_748977|202 .................... .................... .................... .................... .................... ....................
AA 8795 hCoV-19/Denmark/DCGC-19479/2020|EPI_ISL_748978|202 .................... .................... .................... .................... .................... ....................
AA 8796 hCoV-19/Denmark/DCGC-19480/2020|EPI_ISL_748979|202 .................... .................... .................... .................... .................... ....................
AA 8797 hCoV-19/Denmark/DCGC-19482/2020|EPI_ISL_748980|202 .................... .A.................. .................... .................... .................... ...A................
AA 8798 hCoV-19/Denmark/DCGC-19485/2020|EPI_ISL_748981|202 .................... .................... .................... .................... .................... ....................
AA 8799 hCoV-19/Denmark/DCGC-19487/2020|EPI_ISL_748982|202 .................... .................... .................... .................... .................... ....................
AA 8800 hCoV-19/Denmark/DCGC-19488/2020|EPI_ISL_748983|202 .................... .................... .................... .................... .................... ....................
AA 8801 hCoV-19/Denmark/DCGC-19489/2020|EPI_ISL_748984|202 .................... .................... .................... .................... .................... ....................
AA 8802 hCoV-19/Denmark/DCGC-19490/2020|EPI_ISL_748985|202 .................... .................... .................... .................... .................... ....................
AA 8803 hCoV-19/Denmark/DCGC-19492/2020|EPI_ISL_748986|202 .................... .................... .................... .................... .................... ....................
AA 8804 hCoV-19/Denmark/DCGC-19495/2020|EPI_ISL_748987|202 .................... .................... .................... .................... .................... ....................
AA 8805 hCoV-19/Denmark/DCGC-19497/2020|EPI_ISL_748988|202 .................... .................... .................... .................... .................... ....................
AA 8806 hCoV-19/Denmark/DCGC-19498/2020|EPI_ISL_748989|202 .................... .................... .................... .................... .................... ....................
AA 8807 hCoV-19/Denmark/DCGC-19499/2020|EPI_ISL_748990|202 .................... .................... .................... .................... .................... ....................
AA 8808 hCoV-19/Denmark/DCGC-19501/2020|EPI_ISL_748991|202 .................... .................... .................... .................... .................... ....................
AA 8809 hCoV-19/Denmark/DCGC-19502/2020|EPI_ISL_748992|202 .................... .................... .................... .................... .................... ....................
AA 8810 hCoV-19/Denmark/DCGC-19504/2020|EPI_ISL_748993|202 .................... .................... .................... .................... .................... ....................
AA 8811 hCoV-19/Denmark/DCGC-19505/2020|EPI_ISL_748994|202 .................... .................... .................... .................... .................... ....................
AA 8812 hCoV-19/Denmark/DCGC-19507/2020|EPI_ISL_748995|202 .................... .................... .................... .................... .................... ....................
AA 8813 hCoV-19/Denmark/DCGC-19508/2020|EPI_ISL_748996|202 .................... .................... .................... .................... .................... ....................
AA 8814 hCoV-19/Denmark/DCGC-19510/2020|EPI_ISL_748997|202 .................... .................... .................... .................... .................... ....................
AA 8815 hCoV-19/Denmark/DCGC-19512/2020|EPI_ISL_748998|202 .................... .................... .................... .................... .................... ....................
AA 8816 hCoV-19/Denmark/DCGC-19513/2020|EPI_ISL_748999|202 .................... .................... .................... .................... .................... ....................
AA 8817 hCoV-19/USA/CA-CHLA-PLM97595757/2020|EPI_ISL_75349 .................... .A.................. .................... .................... .................... ....................
AA 8818 hCoV-19/USA/CA-CHLA-PLM68468625/2020|EPI_ISL_75349 .................... .................... .................... .................... .................... ....................
AA 8819 hCoV-19/England/MILK-C7AE29/2020|EPI_ISL_757001|20 .................... .........A.......... .................... .................... .................... ....................
AA 8820 hCoV-19/England/QEUH-C7D08A/2020|EPI_ISL_757007|20 .................... .................... .................... .................... .................... ....................
AA 8821 hCoV-19/England/MILK-C78A54/2020|EPI_ISL_757010|20 .................... .................... .................... .................... .................... ....................
AA 8822 hCoV-19/England/QEUH-C7FF4F/2020|EPI_ISL_757011|20 .................... .................... .................... .................... .................... ....................
AA 8823 hCoV-19/England/QEUH-C7C267/2020|EPI_ISL_757012|20 .................... .................... .................... .................... .................... ....................
AA 8824 hCoV-19/England/MILK-C7AE1A/2020|EPI_ISL_757013|20 .................... .................... .................... .................... .................... ....................
AA 8825 hCoV-19/England/MILK-C7B769/2020|EPI_ISL_757014|20 .................... .................... .................... .................... .................... ....................
AA 8826 hCoV-19/Scotland/QEUH-C54371/2020|EPI_ISL_757060|2 .................... .................... .................... .................... .................... ....................
AA 8827 hCoV-19/Scotland/QEUH-C54380/2020|EPI_ISL_757069|2 .................... .................... .................... .................... .................... ....................
AA 8828 hCoV-19/Denmark/DCGC-21671/2020|EPI_ISL_758300|202 .................... .................... .................... .................... .................... ....................
AA 8829 hCoV-19/Denmark/DCGC-21673/2020|EPI_ISL_758301|202 .................... .................... .................... .................... .................... ....................

| | | | | | | | | |
|---|---|---|---|---|---|---|---|---|
| AA | 8830 | hCoV-19/Denmark/DCGC-21676/2020\|EPI_ISL_758302\|202 | ···················· | ···················· | ···················· | ···················· | ···················· | ···················· |
| AA | 8831 | hCoV-19/Denmark/DCGC-21679/2020\|EPI_ISL_758303\|202 | ···················· | ···················· | ···················· | ···················· | ···················· | ···················· |
| AA | 8832 | hCoV-19/Denmark/DCGC-21681/2020\|EPI_ISL_758304\|202 | ···················· | ···················· | ···················· | ···················· | ···················· | ···················· |
| AA | 8833 | hCoV-19/Denmark/DCGC-21682/2020\|EPI_ISL_758305\|202 | ···················· | ···················· | ···················· | ···················· | ···················· | ···················· |
| AA | 8834 | hCoV-19/Denmark/DCGC-21683/2020\|EPI_ISL_758306\|202 | ···················· | ···················· | ···················· | ···················· | ···················· | ···················· |
| AA | 8835 | hCoV-19/Denmark/DCGC-21684/2020\|EPI_ISL_758307\|202 | ···················· | ···················· | ···················· | ···················· | ···················· | ···················· |
| AA | 8836 | hCoV-19/Denmark/DCGC-21685/2020\|EPI_ISL_758308\|202 | ···················· | ···················· | ···················· | ···················· | ···················· | ···················· |
| AA | 8837 | hCoV-19/Denmark/DCGC-21687/2020\|EPI_ISL_758309\|202 | ···················· | ···················· | ···················· | ···················· | ···················· | ···················· |
| AA | 8838 | hCoV-19/Denmark/DCGC-21688/2020\|EPI_ISL_758310\|202 | ···················· | ···················· | ···················· | ···················· | ···················· | ···················· |
| AA | 8839 | hCoV-19/Denmark/DCGC-21689/2020\|EPI_ISL_758311\|202 | ···················· | ···················· | ···················· | ···················· | ···················· | ···················· |
| AA | 8840 | hCoV-19/Denmark/DCGC-21691/2020\|EPI_ISL_758312\|202 | ···················· | ···················· | ···················· | ···················· | ···················· | ···················· |
| AA | 8841 | hCoV-19/Denmark/DCGC-21694/2020\|EPI_ISL_758313\|202 | ···················· | ···················· | ···················· | ···················· | ···················· | ···················· |
| AA | 8842 | hCoV-19/Denmark/DCGC-21695/2020\|EPI_ISL_758314\|202 | ···················· | ···················· | ···················· | ···················· | ···················· | ···················· |
| AA | 8843 | hCoV-19/Denmark/DCGC-21696/2020\|EPI_ISL_758315\|202 | ···················· | ···················· | ···················· | ···················· | ···················· | ···················· |
| AA | 8844 | hCoV-19/Denmark/DCGC-21700/2020\|EPI_ISL_758316\|202 | ···················· | ···················· | ···················· | ···················· | ···················· | ···················· |
| AA | 8845 | hCoV-19/Denmark/DCGC-21701/2020\|EPI_ISL_758317\|202 | ···················· | ···················· | ···················· | ···················· | ···················· | ···················· |
| AA | 8846 | hCoV-19/Denmark/DCGC-21702/2020\|EPI_ISL_758318\|202 | ···················· | ···················· | ···················· | ···················· | ···················· | ···················· |
| AA | 8847 | hCoV-19/Denmark/DCGC-21706/2020\|EPI_ISL_758319\|202 | ···················· | ···················· | ···················· | ···················· | ···················· | ···················· |
| AA | 8848 | hCoV-19/Denmark/DCGC-21707/2020\|EPI_ISL_758320\|202 | ···················· | ·A·················· | ···················· | ···················· | ···················· | ···A················ |
| AA | 8849 | hCoV-19/Denmark/DCGC-21708/2020\|EPI_ISL_758321\|202 | ···················· | ···················· | ···················· | ···················· | ···················· | ···················· |
| AA | 8850 | hCoV-19/Denmark/DCGC-21710/2020\|EPI_ISL_758322\|202 | ···················· | ···················· | ···················· | ···················· | ···················· | ···················· |
| AA | 8851 | hCoV-19/Denmark/DCGC-21711/2020\|EPI_ISL_758323\|202 | ···················· | ···················· | ···················· | ···················· | ···················· | ···················· |
| AA | 8852 | hCoV-19/Denmark/DCGC-21716/2020\|EPI_ISL_758324\|202 | ···················· | ···················· | ···················· | ···················· | ···················· | ···················· |
| AA | 8853 | hCoV-19/Denmark/DCGC-21718/2020\|EPI_ISL_758325\|202 | ···················· | ···················· | ···················· | ···················· | ···················· | ···················· |
| AA | 8854 | hCoV-19/Denmark/DCGC-21720/2020\|EPI_ISL_758326\|202 | ···················· | ···················· | ···················· | ···················· | ···················· | ···················· |
| AA | 8855 | hCoV-19/Denmark/DCGC-21722/2020\|EPI_ISL_758327\|202 | ···················· | ···················· | ···················· | ···················· | ···················· | ···················· |
| AA | 8856 | hCoV-19/Denmark/DCGC-21724/2020\|EPI_ISL_758328\|202 | ···················· | ···················· | ···················· | ···················· | ···················· | ···················· |
| AA | 8857 | hCoV-19/Denmark/DCGC-21726/2020\|EPI_ISL_758329\|202 | ···················· | ···················· | ···················· | ···················· | ···················· | ···················· |
| AA | 8858 | hCoV-19/Denmark/DCGC-21727/2020\|EPI_ISL_758330\|202 | ···················· | ···················· | ···················· | ···················· | ···················· | ···················· |
| AA | 8859 | hCoV-19/Denmark/DCGC-21729/2020\|EPI_ISL_758331\|202 | ···················· | ···················· | ···T················ | ···················· | ···················· | ···················· |
| AA | 8860 | hCoV-19/Denmark/DCGC-21730/2020\|EPI_ISL_758332\|202 | ···················· | ···················· | ···················· | ···················· | ···················· | ···················· |
| AA | 8861 | hCoV-19/Denmark/DCGC-21733/2020\|EPI_ISL_758333\|202 | ···················· | ···················· | ···················· | ···················· | ···················· | ···················· |
| AA | 8862 | hCoV-19/Denmark/DCGC-21734/2020\|EPI_ISL_758334\|202 | ···················· | ···················· | ···················· | ···················· | ···················· | ···················· |
| AA | 8863 | hCoV-19/Denmark/DCGC-21735/2020\|EPI_ISL_758335\|202 | ···················· | ···················· | ···················· | ···················· | ···················· | ···················· |
| AA | 8864 | hCoV-19/Denmark/DCGC-21736/2020\|EPI_ISL_758336\|202 | ···················· | ···················· | ···················· | ···················· | ···················· | ···················· |
| AA | 8865 | hCoV-19/Denmark/DCGC-21739/2020\|EPI_ISL_758337\|202 | ···················· | ···················· | ···················· | ···················· | ···················· | ···················· |
| AA | 8866 | hCoV-19/Denmark/DCGC-21740/2020\|EPI_ISL_758338\|202 | ···················· | ···················· | ···················· | ···················· | ···················· | ···················· |
| AA | 8867 | hCoV-19/Denmark/DCGC-21741/2020\|EPI_ISL_758339\|202 | ···················· | ···················· | ···················· | ···················· | ···················· | ···················· |
| AA | 8868 | hCoV-19/Denmark/DCGC-21744/2020\|EPI_ISL_758340\|202 | ···················· | ···················· | ···················· | ···················· | ···················· | ···················· |
| AA | 8869 | hCoV-19/Denmark/DCGC-21747/2020\|EPI_ISL_758341\|202 | ···················· | ···················· | ···················· | ···················· | ···················· | ···················· |
| AA | 8870 | hCoV-19/Denmark/DCGC-21750/2020\|EPI_ISL_758342\|202 | ···················· | ···················· | ···················· | ···················· | ···················· | ···················· |
| AA | 8871 | hCoV-19/Denmark/DCGC-21752/2020\|EPI_ISL_758343\|202 | ···················· | ···················· | ···················· | ···················· | ···················· | ···················· |
| AA | 8872 | hCoV-19/Denmark/DCGC-21755/2020\|EPI_ISL_758344\|202 | ···················· | ···················· | ···················· | ···················· | ···················· | ···················· |
| AA | 8873 | hCoV-19/Denmark/DCGC-21756/2020\|EPI_ISL_758345\|202 | ···················· | ···················· | ···················· | ···················· | ···················· | ···················· |
| AA | 8874 | hCoV-19/Denmark/DCGC-21757/2020\|EPI_ISL_758346\|202 | ···················· | ···················· | ···················· | ···················· | ···················· | ···················· |
| AA | 8875 | hCoV-19/Denmark/DCGC-21758/2020\|EPI_ISL_758347\|202 | ···················· | ···················· | ···················· | ···················· | ···················· | ···················· |
| AA | 8876 | hCoV-19/Denmark/DCGC-21759/2020\|EPI_ISL_758348\|202 | ···················· | ···················· | ···················· | ···················· | ···················· | ···················· |
| AA | 8877 | hCoV-19/Denmark/DCGC-21762/2020\|EPI_ISL_758349\|202 | ···················· | ···················· | ···················· | ···················· | ···················· | ···················· |
| AA | 8878 | hCoV-19/Denmark/DCGC-21829/2020\|EPI_ISL_758390\|202 | ···················· | ···················· | ···················· | ···················· | ···················· | ···················· |
| AA | 8879 | hCoV-19/Denmark/DCGC-21831/2020\|EPI_ISL_758391\|202 | ···················· | ···················· | ···················· | ···················· | ···················· | ···················· |
| AA | 8880 | hCoV-19/Denmark/DCGC-21835/2020\|EPI_ISL_758394\|202 | ···················· | ···················· | ···················· | ···················· | ···················· | ···················· |
| AA | 8881 | hCoV-19/Denmark/DCGC-21184/2020\|EPI_ISL_759600\|202 | ···················· | ···················· | ···················· | ···················· | ···················· | ···················· |
| AA | 8882 | hCoV-19/Denmark/DCGC-21185/2020\|EPI_ISL_759601\|202 | ···················· | ···················· | ···················· | ···················· | ···················· | ···················· |
| AA | 8883 | hCoV-19/Denmark/DCGC-21186/2020\|EPI_ISL_759602\|202 | ···················· | ···················· | ···················· | ···················· | ···················· | ···················· |
| AA | 8884 | hCoV-19/Denmark/DCGC-21187/2020\|EPI_ISL_759603\|202 | ···················· | ···················· | ···················· | ···················· | ···················· | ···················· |
| AA | 8885 | hCoV-19/Denmark/DCGC-21188/2020\|EPI_ISL_759604\|202 | ···················· | ···················· | ···················· | ···················· | ···················· | ···················· |
| AA | 8886 | hCoV-19/Denmark/DCGC-21191/2020\|EPI_ISL_759605\|202 | ···················· | ···················· | ···················· | ···················· | ···················· | ···················· |
| AA | 8887 | hCoV-19/Denmark/DCGC-21192/2020\|EPI_ISL_759606\|202 | ···················· | ···················· | ···················· | ···················· | ···················· | ···················· |
| AA | 8888 | hCoV-19/Denmark/DCGC-21193/2020\|EPI_ISL_759607\|202 | ···················· | ···················· | ···················· | ···················· | ···················· | ···················· |
| AA | 8889 | hCoV-19/Denmark/DCGC-21195/2020\|EPI_ISL_759608\|202 | ···················· | ···················· | ···················· | ···················· | ···················· | ···················· |
| AA | 8890 | hCoV-19/Denmark/DCGC-21197/2020\|EPI_ISL_759609\|202 | ···················· | ···················· | ···················· | ···················· | ···················· | ···················· |
| AA | 8891 | hCoV-19/Denmark/DCGC-21201/2020\|EPI_ISL_759610\|202 | ···················· | ···················· | ···················· | ···················· | ···················· | ···············A···· |
| AA | 8892 | hCoV-19/Denmark/DCGC-21202/2020\|EPI_ISL_759611\|202 | ···················· | ···················· | ···················· | ···················· | ···················· | ···················· |
| AA | 8893 | hCoV-19/Denmark/DCGC-21207/2020\|EPI_ISL_759612\|202 | ···················· | ···················· | ···················· | ···················· | ···················· | ···················· |
| AA | 8894 | hCoV-19/Denmark/DCGC-21210/2020\|EPI_ISL_759613\|202 | ···················· | ···················· | ···················· | ···················· | ···················· | ···················· |
| AA | 8895 | hCoV-19/Denmark/DCGC-21211/2020\|EPI_ISL_759614\|202 | ···················· | ···················· | ···················· | ···················· | ···················· | ···················· |
| AA | 8896 | hCoV-19/Denmark/DCGC-21216/2020\|EPI_ISL_759615\|202 | ···················· | ···················· | ···················· | ···················· | ···················· | ···················· |
| AA | 8897 | hCoV-19/Denmark/DCGC-21217/2020\|EPI_ISL_759616\|202 | ···················· | ···················· | ···················· | ···················· | ···················· | ···················· |
| AA | 8898 | hCoV-19/Denmark/DCGC-21220/2020\|EPI_ISL_759617\|202 | ···················· | ···················· | ···················· | ···················· | ···················· | ···················· |
| AA | 8899 | hCoV-19/Denmark/DCGC-21221/2020\|EPI_ISL_759618\|202 | ···················· | ···················· | ···················· | ···················· | ···················· | ···················· |
| AA | 8900 | hCoV-19/Denmark/DCGC-21223/2020\|EPI_ISL_759619\|202 | ···················· | ···················· | ···················· | ···················· | ···················· | ···················· |
| AA | 8901 | hCoV-19/Denmark/DCGC-21225/2020\|EPI_ISL_759620\|202 | ···················· | ···················· | ···················· | ···················· | ···················· | ···················· |
| AA | 8902 | hCoV-19/Denmark/DCGC-21227/2020\|EPI_ISL_759621\|202 | ···················· | ···················· | ···················· | ···················· | ···················· | ···················· |
| AA | 8903 | hCoV-19/Denmark/DCGC-21228/2020\|EPI_ISL_759622\|202 | ···················· | ···················· | ···················· | ···················· | ···················· | ···················· |
| AA | 8904 | hCoV-19/Denmark/DCGC-21230/2020\|EPI_ISL_759623\|202 | ···················· | ···················· | ···················· | ···················· | ···················· | ···················· |
| AA | 8905 | hCoV-19/Denmark/DCGC-21231/2020\|EPI_ISL_759624\|202 | ···················· | ···················· | ···················· | ···················· | ···················· | ···················· |
| AA | 8906 | hCoV-19/Denmark/DCGC-21232/2020\|EPI_ISL_759625\|202 | ···················· | ···················· | ···················· | ···················· | ···················· | ···················· |
| AA | 8907 | hCoV-19/Denmark/DCGC-21234/2020\|EPI_ISL_759626\|202 | ···················· | ···················· | ···················· | ···················· | ···················· | ···················· |
| AA | 8908 | hCoV-19/Denmark/DCGC-21236/2020\|EPI_ISL_759627\|202 | ···················· | ···················· | ···················· | ···················· | ···················· | ···················· |
| AA | 8909 | hCoV-19/Denmark/DCGC-21237/2020\|EPI_ISL_759628\|202 | ···················· | ···················· | ···················· | ···················· | ···················· | ···················· |

| | | | | | | | | |
|---|---|---|---|---|---|---|---|---|
| AA | 8910 | hCoV-19/Denmark/DCGC-21239/2020\|EPI_ISL_759629\|202 | ···················· | ···················· | ···················· | ···················· | ···················· | ···················· |
| AA | 8911 | hCoV-19/Denmark/DCGC-21241/2020\|EPI_ISL_759630\|202 | ···················· | ···················· | ···················· | ···················· | ···················· | ···················· |
| AA | 8912 | hCoV-19/Denmark/DCGC-21243/2020\|EPI_ISL_759631\|202 | ···················· | ···················· | ···················· | ···················· | ···················· | ···················· |
| AA | 8913 | hCoV-19/Denmark/DCGC-21244/2020\|EPI_ISL_759632\|202 | ···················· | ···················· | ···················· | ···················· | ···················· | ···················· |
| AA | 8914 | hCoV-19/Denmark/DCGC-21245/2020\|EPI_ISL_759633\|202 | ···················· | ···················· | ···················· | ···················· | ···················· | ···················· |
| AA | 8915 | hCoV-19/Denmark/DCGC-21251/2020\|EPI_ISL_759634\|202 | ···················· | ···················· | ···················· | ···················· | ···················· | ···················· |
| AA | 8916 | hCoV-19/Denmark/DCGC-21252/2020\|EPI_ISL_759635\|202 | ···················· | ···················· | ···················· | ···················· | ···················· | ···················· |
| AA | 8917 | hCoV-19/Denmark/DCGC-21253/2020\|EPI_ISL_759636\|202 | ···················· | ···················· | ···················· | ···················· | ···················· | ···················· |
| AA | 8918 | hCoV-19/Denmark/DCGC-21255/2020\|EPI_ISL_759637\|202 | ···················· | ···················· | ···················· | ···················· | ···················· | ···················· |
| AA | 8919 | hCoV-19/Denmark/DCGC-21256/2020\|EPI_ISL_759638\|202 | ···················· | ···················· | ···················· | ···················· | ···················· | ···················· |
| AA | 8920 | hCoV-19/Denmark/DCGC-21257/2020\|EPI_ISL_759639\|202 | ···················· | ···················· | ···················· | ···················· | ···················· | ···················· |
| AA | 8921 | hCoV-19/Denmark/DCGC-21262/2020\|EPI_ISL_759640\|202 | ···················· | ···················· | ···················· | ···················· | ···················· | ···················· |
| AA | 8922 | hCoV-19/Denmark/DCGC-21264/2020\|EPI_ISL_759641\|202 | ···················· | ···················· | ···················· | ···················· | ···················· | ···················· |
| AA | 8923 | hCoV-19/Denmark/DCGC-21265/2020\|EPI_ISL_759642\|202 | ···················· | ···················· | ···················· | ···················· | ···················· | ···················· |
| AA | 8924 | hCoV-19/Denmark/DCGC-21266/2020\|EPI_ISL_759643\|202 | ···················· | ···················· | ···················· | ···················· | ···················· | ···················· |
| AA | 8925 | hCoV-19/Denmark/DCGC-21268/2020\|EPI_ISL_759644\|202 | ···················· | ···················· | ···················· | ···················· | ···················· | ···················· |
| AA | 8926 | hCoV-19/Denmark/DCGC-21269/2020\|EPI_ISL_759645\|202 | ···················· | ···················· | ···················· | ···················· | ···················· | ···················· |
| AA | 8927 | hCoV-19/Denmark/DCGC-21271/2020\|EPI_ISL_759646\|202 | ···················· | ···················· | ···················· | ···················· | ···················· | ···················· |
| AA | 8928 | hCoV-19/Denmark/DCGC-21272/2020\|EPI_ISL_759647\|202 | ···················· | ···················· | ···················· | ···················· | ···················· | ···················· |
| AA | 8929 | hCoV-19/Denmark/DCGC-21273/2020\|EPI_ISL_759648\|202 | ···················· | ···················· | ···················· | ···················· | ···················· | ···················· |
| AA | 8930 | hCoV-19/Denmark/DCGC-21274/2020\|EPI_ISL_759649\|202 | ···················· | ·A·················· | ···················· | ···················· | ···················· | ···················· |
| AA | 8931 | hCoV-19/Denmark/DCGC-21275/2020\|EPI_ISL_759650\|202 | ···················· | ···················· | ···················· | ···················· | ···················· | ···················· |
| AA | 8932 | hCoV-19/Denmark/DCGC-21281/2020\|EPI_ISL_759651\|202 | ···················· | ···················· | ···················· | ···················· | ···················· | ···················· |
| AA | 8933 | hCoV-19/Denmark/DCGC-21282/2020\|EPI_ISL_759652\|202 | ···················· | ···················· | ···················· | ···················· | ···················· | ···················· |
| AA | 8934 | hCoV-19/Denmark/DCGC-21283/2020\|EPI_ISL_759653\|202 | ···················· | ···················· | ···················· | ···················· | ···················· | ···················· |
| AA | 8935 | hCoV-19/Denmark/DCGC-21285/2020\|EPI_ISL_759654\|202 | ···················· | ···················· | ···················· | ···················· | ···················· | ···················· |
| AA | 8936 | hCoV-19/Denmark/DCGC-21286/2020\|EPI_ISL_759655\|202 | ···················· | ···················· | ···················· | ···················· | ···················· | ···················· |
| AA | 8937 | hCoV-19/Denmark/DCGC-21289/2020\|EPI_ISL_759656\|202 | ···················· | ···················· | ···················· | ···················· | ···················· | ···················· |
| AA | 8938 | hCoV-19/Denmark/DCGC-21293/2020\|EPI_ISL_759657\|202 | ···················· | ···················· | ···················· | ···················· | ···················· | ···················· |
| AA | 8939 | hCoV-19/Denmark/DCGC-21296/2020\|EPI_ISL_759658\|202 | ···················· | ···················· | ···················· | ···················· | ···················· | ···················· |
| AA | 8940 | hCoV-19/Denmark/DCGC-21299/2020\|EPI_ISL_759659\|202 | ···················· | ·A·················· | ···················· | ···················· | ···················· | ··A················· |
| AA | 8941 | hCoV-19/Denmark/DCGC-21300/2020\|EPI_ISL_759660\|202 | ···················· | ···················· | ···················· | ···················· | ···················· | ···················· |
| AA | 8942 | hCoV-19/Denmark/DCGC-21302/2020\|EPI_ISL_759661\|202 | ···················· | ···················· | ···················· | ···················· | ···················· | ···················· |
| AA | 8943 | hCoV-19/Denmark/DCGC-21303/2020\|EPI_ISL_759662\|202 | ···················· | ·A·················· | ···················· | ···················· | ···················· | ···················· |
| AA | 8944 | hCoV-19/Denmark/DCGC-21304/2020\|EPI_ISL_759663\|202 | ···················· | ···················· | ···················· | ···················· | ···················· | ···················· |
| AA | 8945 | hCoV-19/Denmark/DCGC-21306/2020\|EPI_ISL_759664\|202 | ···················· | ···················· | ···················· | ···················· | ···················· | ···················· |
| AA | 8946 | hCoV-19/Denmark/DCGC-21307/2020\|EPI_ISL_759665\|202 | ···················· | ···················· | ···················· | ···················· | ···················· | ···················· |
| AA | 8947 | hCoV-19/Denmark/DCGC-21308/2020\|EPI_ISL_759666\|202 | ···················· | ···················· | ···················· | ···················· | ···················· | ···················· |
| AA | 8948 | hCoV-19/Denmark/DCGC-21310/2020\|EPI_ISL_759667\|202 | ···················· | ···················· | ···················· | ···················· | ···················· | ···················· |
| AA | 8949 | hCoV-19/Denmark/DCGC-21312/2020\|EPI_ISL_759668\|202 | ···················· | ···················· | ···················· | ···················· | ···················· | ···················· |
| AA | 8950 | hCoV-19/Denmark/DCGC-21314/2020\|EPI_ISL_759669\|202 | ···················· | ···················· | ···················· | ···················· | ···················· | ···················· |
| AA | 8951 | hCoV-19/Denmark/DCGC-21315/2020\|EPI_ISL_759670\|202 | ···················· | ···················· | ···················· | ···················· | ···················· | ···················· |
| AA | 8952 | hCoV-19/Denmark/DCGC-21316/2020\|EPI_ISL_759671\|202 | ···················· | ···················· | ···················· | ···················· | ···················· | ···················· |
| AA | 8953 | hCoV-19/Denmark/DCGC-21317/2020\|EPI_ISL_759672\|202 | ···················· | ···················· | ···················· | ···················· | ···················· | ···················· |
| AA | 8954 | hCoV-19/Denmark/DCGC-21319/2020\|EPI_ISL_759673\|202 | ···················· | ···················· | ···················· | ···················· | ···················· | ···················· |
| AA | 8955 | hCoV-19/Denmark/DCGC-21320/2020\|EPI_ISL_759674\|202 | ···················· | ···················· | ···················· | ···················· | ···················· | ···················· |
| AA | 8956 | hCoV-19/Denmark/DCGC-21321/2020\|EPI_ISL_759675\|202 | ···················· | ···················· | ···················· | ···················· | ···················· | ···················· |
| AA | 8957 | hCoV-19/Denmark/DCGC-21322/2020\|EPI_ISL_759676\|202 | ···················· | ···················· | ···················· | ···················· | ···················· | ···················· |
| AA | 8958 | hCoV-19/Denmark/DCGC-21324/2020\|EPI_ISL_759677\|202 | ···················· | ···················· | ···················· | ···················· | ···················· | ···················· |
| AA | 8959 | hCoV-19/Denmark/DCGC-21325/2020\|EPI_ISL_759678\|202 | ···················· | ···················· | ···················· | ···················· | ···················· | ···················· |
| AA | 8960 | hCoV-19/Denmark/DCGC-21326/2020\|EPI_ISL_759679\|202 | ···················· | ···················· | ···················· | ···················· | ···················· | ···················· |
| AA | 8961 | hCoV-19/USA/NY-Wadsworth-251094-01/2020\|EPI_ISL_75 | ···················· | ···················· | ···················· | ···················· | ···················· | ···················· |
| AA | 8962 | hCoV-19/USA/NY-Wadsworth-250906-01/2020\|EPI_ISL_75 | ···················· | ···················· | ···················· | ···················· | ···················· | ···················· |
| AA | 8963 | hCoV-19/USA/NY-Wadsworth-250903-01/2020\|EPI_ISL_75 | ···················· | ···················· | ···················· | ···················· | ···················· | ···················· |
| AA | 8964 | hCoV-19/USA/NY-Wadsworth-250902-01/2020\|EPI_ISL_75 | ···················· | ···················· | ···················· | ···················· | ···················· | ···················· |
| AA | 8965 | hCoV-19/USA/NY-Wadsworth-251079-01/2020\|EPI_ISL_75 | ···················· | ···················· | ···················· | ···················· | ···················· | ···················· |
| AA | 8966 | hCoV-19/USA/NY-Wadsworth-251076-01/2020\|EPI_ISL_75 | ···················· | ···················· | ···················· | ···················· | ···················· | ···················· |
| AA | 8967 | hCoV-19/USA/NY-Wadsworth-251071-01/2020\|EPI_ISL_75 | ···················· | ···················· | ···················· | ···················· | ···················· | ···················· |
| AA | 8968 | hCoV-19/England/QEUH-C7C276/2020\|EPI_ISL_757021\|20 | ···················· | ···················· | ···················· | ···················· | ······T············· | ···················· |
| AA | 8969 | hCoV-19/England/QEUH-C7D07B/2020\|EPI_ISL_757028\|20 | ···················· | ···················· | ···················· | ···················· | ···················· | ···················· |
| AA | 8970 | hCoV-19/England/QEUH-C7FF21/2020\|EPI_ISL_757032\|20 | ···················· | ···················· | ···················· | ···················· | ···················· | ···················· |
| AA | 8971 | hCoV-19/Scotland/QEUH-C5B86C/2020\|EPI_ISL_757034\|2 | ···················· | ···················· | ···················· | ···················· | ···················· | ···················· |
| AA | 8972 | hCoV-19/Scotland/QEUH-C5B8C6/2020\|EPI_ISL_757035\|2 | ···················· | ···················· | ···················· | ···················· | ···················· | ···················· |
| AA | 8973 | hCoV-19/Scotland/QEUH-C5CB35/2020\|EPI_ISL_757036\|2 | ···················· | ···················· | ···················· | ···················· | ···················· | ···················· |
| AA | 8974 | hCoV-19/Scotland/QEUH-C5C76E/2020\|EPI_ISL_757037\|2 | ···················· | ···················· | ···················· | ···················· | ···················· | ···················· |
| AA | 8975 | hCoV-19/Scotland/QEUH-C5B820/2020\|EPI_ISL_757040\|2 | ···················· | ···················· | ···················· | ···················· | ···················· | ···················· |
| AA | 8976 | hCoV-19/Scotland/QEUH-C5C77D/2020\|EPI_ISL_757041\|2 | ···················· | ···················· | ···················· | ···················· | ···················· | ···················· |
| AA | 8977 | hCoV-19/Scotland/QEUH-C54362/2020\|EPI_ISL_757042\|2 | ···················· | ···················· | ···················· | ···················· | ···················· | ···················· |
| AA | 8978 | hCoV-19/Scotland/QEUH-C543BD/2020\|EPI_ISL_757043\|2 | ···················· | ···················· | ···················· | ···················· | ···················· | ···················· |
| AA | 8979 | hCoV-19/Scotland/QEUH-C54423/2020\|EPI_ISL_757044\|2 | ···················· | ···················· | ···················· | ···················· | ···················· | ···················· |
| AA | 8980 | hCoV-19/Scotland/QEUH-C5447E/2020\|EPI_ISL_757045\|2 | ···················· | ···················· | ···················· | ···················· | ···················· | ···················· |
| AA | 8981 | hCoV-19/Scotland/QEUH-C54450/2020\|EPI_ISL_757046\|2 | ···················· | ···················· | ···················· | ···················· | ···················· | ···················· |
| AA | 8982 | hCoV-19/Scotland/QEUH-C540E3/2020\|EPI_ISL_757047\|2 | ···················· | ···················· | ···················· | ···················· | ···················· | ···················· |
| AA | 8983 | hCoV-19/Scotland/QEUH-C543EA/2020\|EPI_ISL_757048\|2 | ···················· | ···················· | ···················· | ···················· | ···················· | ···················· |
| AA | 8984 | hCoV-19/Scotland/QEUH-C543CC/2020\|EPI_ISL_757049\|2 | ···················· | ···················· | ···················· | ···················· | ···················· | ···················· |
| AA | 8985 | hCoV-19/Scotland/QEUH-C5455D/2020\|EPI_ISL_757050\|2 | ···················· | ···················· | ···················· | ···················· | ···················· | ···················· |
| AA | 8986 | hCoV-19/Scotland/QEUH-C543AE/2020\|EPI_ISL_757051\|2 | ···················· | ···················· | ···················· | ···················· | ···················· | ···················· |
| AA | 8987 | hCoV-19/Scotland/QEUH-C54326/2020\|EPI_ISL_757052\|2 | ···················· | ···················· | ···················· | ···················· | ···················· | ···················· |
| AA | 8988 | hCoV-19/Scotland/QEUH-C543DB/2020\|EPI_ISL_757053\|2 | ···················· | ···················· | ···················· | ···················· | ···················· | ···················· |
| AA | 8989 | hCoV-19/Scotland/QEUH-C542ED/2020\|EPI_ISL_757055\|2 | ···················· | ···················· | ···················· | ···················· | ···················· | ···················· |

| | | | | | | | |
|---|---|---|---|---|---|---|---|
| AA | 8990 hCoV-19/Scotland/QEUH-C540C5/2020\|EPI_ISL_757058\|2 | .................... | .................... | ..................... | .................... | .................... | .................... |
| AA | 8991 hCoV-19/Scotland/QEUH-C541B3/2020\|EPI_ISL_757059\|2 | .................... | .................... | ..................... | .................... | .................... | .................... |
| AA | 8992 hCoV-19/Scotland/QEUH-C54159/2020\|EPI_ISL_757061\|2 | .................... | .................... | ..................... | .................... | .................... | .................... |
| AA | 8993 hCoV-19/Scotland/QEUH-C5412C/2020\|EPI_ISL_757062\|2 | .................... | .................... | ..................... | .................... | .................... | .................... |
| AA | 8994 hCoV-19/Scotland/QEUH-C54344/2020\|EPI_ISL_757063\|2 | .................... | .................... | ..................... | .................... | .................... | .................... |
| AA | 8995 hCoV-19/Scotland/QEUH-C5448D/2020\|EPI_ISL_757064\|2 | .................... | .................... | ..................... | .................... | .................... | .................... |
| AA | 8996 hCoV-19/Scotland/QEUH-C54432/2020\|EPI_ISL_757065\|2 | .................... | .................... | ..................... | .................... | .................... | .................... |
| AA | 8997 hCoV-19/Scotland/QEUH-C54405/2020\|EPI_ISL_757066\|2 | .................... | .................... | ..................... | .................... | .................... | .................... |
| AA | 8998 hCoV-19/Scotland/QEUH-C543F9/2020\|EPI_ISL_757067\|2 | .................... | .................... | ..................... | .................... | .................... | .................... |
| AA | 8999 hCoV-19/Scotland/QEUH-C540D4/2020\|EPI_ISL_757068\|2 | .................... | .................... | ..................... | .................... | .................... | .................... |
| AA | 9000 hCoV-19/Scotland/QEUH-C542FC/2020\|EPI_ISL_757070\|2 | .................... | .................... | ..................... | .................... | .................... | .................... |
| AA | 9001 hCoV-19/Scotland/QEUH-C54317/2020\|EPI_ISL_757073\|2 | .................... | .................... | ..................... | .................... | .................... | .................... |
| AA | 9002 hCoV-19/Scotland/QEUH-C542CF/2020\|EPI_ISL_757074\|2 | .................... | .................... | ..................... | .................... | .................... | .................... |
| AA | 9003 hCoV-19/Scotland/QEUH-C54414/2020\|EPI_ISL_757075\|2 | .................... | .................... | ..................... | .................... | .................... | .................... |
| AA | 9004 hCoV-19/Scotland/QEUH-C5413B/2020\|EPI_ISL_757076\|2 | .................... | .................... | ..................... | .................... | .................... | .................... |
| AA | 9005 hCoV-19/Scotland/QEUH-C5439F/2020\|EPI_ISL_757077\|2 | .................... | .................... | ..................... | .................... | .................... | .................... |
| AA | 9006 hCoV-19/Scotland/QEUH-C54308/2020\|EPI_ISL_757079\|2 | .................... | .................... | ..................... | .................... | .................... | .................... |
| AA | 9007 hCoV-19/Scotland/QEUH-C5446F/2020\|EPI_ISL_757080\|2 | .................... | .................... | ..................... | .................... | .................... | .................... |
| AA | 9008 hCoV-19/Scotland/QEUH-C53E44/2020\|EPI_ISL_757081\|2 | .................... | .................... | ..................... | .................... | .....T.............. | .................... |
| AA | 9009 hCoV-19/Scotland/QEUH-C54168/2020\|EPI_ISL_757082\|2 | .................... | .................... | ..................... | .................... | .................... | .................... |
| AA | 9010 hCoV-19/Scotland/QEUH-C4BF1F/2020\|EPI_ISL_757087\|2 | .................... | .................... | ..................... | .................... | .................... | .................... |
| AA | 9011 hCoV-19/Scotland/QEUH-C4BE4F/2020\|EPI_ISL_757088\|2 | .................... | .................... | ..................... | .................... | .................... | .................... |
| AA | 9012 hCoV-19/Scotland/QEUH-C4C02A/2020\|EPI_ISL_757091\|2 | .................... | .................... | ..................... | .................... | .................... | .................... |
| AA | 9013 hCoV-19/Scotland/QEUH-C4BF3D/2020\|EPI_ISL_757096\|2 | .................... | .................... | ..................... | .................... | .................... | .................... |
| AA | 9014 hCoV-19/Scotland/QEUH-C4C1AF/2020\|EPI_ISL_757099\|2 | .................... | .................... | ..................... | .................... | .................... | .................... |
| AA | 9015 hCoV-19/Denmark/DCGC-21765/2020\|EPI_ISL_758350\|202 | .................... | .................... | ..................... | .................... | .................... | .................... |
| AA | 9016 hCoV-19/Denmark/DCGC-21767/2020\|EPI_ISL_758351\|202 | .................... | .................... | ..................... | .................... | .................... | .................... |
| AA | 9017 hCoV-19/Denmark/DCGC-21772/2020\|EPI_ISL_758352\|202 | .................... | .................... | ..................... | .................... | .................... | .................... |
| AA | 9018 hCoV-19/Denmark/DCGC-21773/2020\|EPI_ISL_758353\|202 | .................... | .................... | ..................... | .................... | .................... | .................... |
| AA | 9019 hCoV-19/Denmark/DCGC-21774/2020\|EPI_ISL_758354\|202 | .................... | .................... | ..................... | .................... | .................... | .................... |
| AA | 9020 hCoV-19/Denmark/DCGC-21776/2020\|EPI_ISL_758355\|202 | .................... | .................... | ..................... | .................... | .................... | .................... |
| AA | 9021 hCoV-19/Denmark/DCGC-21778/2020\|EPI_ISL_758356\|202 | .................... | .................... | ..................... | .................... | .................... | .................... |
| AA | 9022 hCoV-19/Denmark/DCGC-21781/2020\|EPI_ISL_758357\|202 | .................... | .................... | ..................... | .................... | .................... | .................... |
| AA | 9023 hCoV-19/Denmark/DCGC-21782/2020\|EPI_ISL_758358\|202 | .................... | .................... | ..................... | .................... | .................... | .................... |
| AA | 9024 hCoV-19/Denmark/DCGC-21783/2020\|EPI_ISL_758359\|202 | .................... | .................... | ..................... | .................... | .................... | .................... |
| AA | 9025 hCoV-19/Denmark/DCGC-21784/2020\|EPI_ISL_758360\|202 | .................... | .................... | ..................... | .................... | .................... | .................... |
| AA | 9026 hCoV-19/Denmark/DCGC-21785/2020\|EPI_ISL_758361\|202 | .................... | .................... | ..................... | .................... | .................... | .................... |
| AA | 9027 hCoV-19/Denmark/DCGC-21787/2020\|EPI_ISL_758362\|202 | .................... | .................... | ..................... | .................... | .................... | .................... |
| AA | 9028 hCoV-19/Denmark/DCGC-21791/2020\|EPI_ISL_758363\|202 | .................... | .................... | ..................... | .................... | .................... | .................... |
| AA | 9029 hCoV-19/Denmark/DCGC-21792/2020\|EPI_ISL_758364\|202 | .................... | .................... | ..................... | .................... | .................... | .................... |
| AA | 9030 hCoV-19/Denmark/DCGC-21794/2020\|EPI_ISL_758365\|202 | .................... | .................... | ..................... | .................... | .................... | .................... |
| AA | 9031 hCoV-19/Denmark/DCGC-21795/2020\|EPI_ISL_758366\|202 | .................... | .................... | ..................... | .................... | .................... | .................... |
| AA | 9032 hCoV-19/Denmark/DCGC-21796/2020\|EPI_ISL_758367\|202 | .................... | .................... | ..................... | .................... | .................... | .................... |
| AA | 9033 hCoV-19/Denmark/DCGC-21797/2020\|EPI_ISL_758368\|202 | .................... | .................... | ..................... | .................... | .................... | .................... |
| AA | 9034 hCoV-19/Denmark/DCGC-21798/2020\|EPI_ISL_758369\|202 | .................... | .................... | ..................... | .................... | .................... | .................... |
| AA | 9035 hCoV-19/Denmark/DCGC-21799/2020\|EPI_ISL_758370\|202 | .................... | .................... | ..................... | .................... | .................... | .................... |
| AA | 9036 hCoV-19/Denmark/DCGC-21801/2020\|EPI_ISL_758371\|202 | .................... | .................... | ..................... | .................... | .................... | .................... |
| AA | 9037 hCoV-19/Denmark/DCGC-21802/2020\|EPI_ISL_758372\|202 | .................... | .................... | ..................... | .................... | .................... | .................... |
| AA | 9038 hCoV-19/Denmark/DCGC-21803/2020\|EPI_ISL_758373\|202 | .................... | .................... | ..................... | .................... | .................... | .................... |
| AA | 9039 hCoV-19/Denmark/DCGC-21804/2020\|EPI_ISL_758374\|202 | .................... | .................... | ..................... | .................... | .................... | .................... |
| AA | 9040 hCoV-19/Denmark/DCGC-21806/2020\|EPI_ISL_758375\|202 | .................... | .................... | ..................... | .................... | .................... | .................... |
| AA | 9041 hCoV-19/Denmark/DCGC-21807/2020\|EPI_ISL_758376\|202 | .................... | .................... | ..................... | .................... | .................... | .................... |
| AA | 9042 hCoV-19/Denmark/DCGC-21808/2020\|EPI_ISL_758377\|202 | .................... | .................... | ..................... | .................... | .................... | .................... |
| AA | 9043 hCoV-19/Denmark/DCGC-21809/2020\|EPI_ISL_758378\|202 | .................... | .................... | ..................... | .................... | .................... | .................... |
| AA | 9044 hCoV-19/Denmark/DCGC-21812/2020\|EPI_ISL_758379\|202 | .................... | .................... | ..................... | .................... | .................... | .................... |
| AA | 9045 hCoV-19/Denmark/DCGC-21813/2020\|EPI_ISL_758380\|202 | .................... | .................... | ..................... | .................... | .................... | .................... |
| AA | 9046 hCoV-19/Denmark/DCGC-21814/2020\|EPI_ISL_758381\|202 | .................... | .................... | ..................... | .................... | .................... | .................... |
| AA | 9047 hCoV-19/Denmark/DCGC-21815/2020\|EPI_ISL_758382\|202 | .................... | .................... | ..................... | .................... | .................... | .................... |
| AA | 9048 hCoV-19/Denmark/DCGC-21817/2020\|EPI_ISL_758383\|202 | .................... | .................... | ..................... | .................... | .................... | .................... |
| AA | 9049 hCoV-19/Denmark/DCGC-21818/2020\|EPI_ISL_758384\|202 | .................... | .................... | ..................... | .................... | .................... | .................... |
| AA | 9050 hCoV-19/Denmark/DCGC-21819/2020\|EPI_ISL_758385\|202 | .................... | .................... | ..................... | .................... | .................... | .................... |
| AA | 9051 hCoV-19/Denmark/DCGC-21821/2020\|EPI_ISL_758386\|202 | .................... | .................... | ..................... | .................... | .................... | .................... |
| AA | 9052 hCoV-19/Denmark/DCGC-21822/2020\|EPI_ISL_758387\|202 | .................... | .A.................. | ..................... | .................... | .................... | ..A................. |
| AA | 9053 hCoV-19/Denmark/DCGC-21823/2020\|EPI_ISL_758388\|202 | .................... | .................... | ..................... | .................... | .................... | .................... |
| AA | 9054 hCoV-19/Denmark/DCGC-21828/2020\|EPI_ISL_758389\|202 | .................... | .................... | ..................... | .................... | .................... | .................... |
| AA | 9055 hCoV-19/Denmark/DCGC-21832/2020\|EPI_ISL_758392\|202 | .................... | .................... | ..................... | .................... | .................... | .................... |
| AA | 9056 hCoV-19/Denmark/DCGC-21834/2020\|EPI_ISL_758393\|202 | .................... | .................... | ..................... | .................... | .................... | .................... |
| AA | 9057 hCoV-19/Denmark/DCGC-21838/2020\|EPI_ISL_758395\|202 | .................... | .................... | ..................... | .................... | .................... | .................... |
| AA | 9058 hCoV-19/Denmark/DCGC-21839/2020\|EPI_ISL_758396\|202 | .................... | .................... | ..................... | .................... | .................... | .................... |
| AA | 9059 hCoV-19/Denmark/DCGC-21840/2020\|EPI_ISL_758397\|202 | .................... | .................... | ..................... | .................... | .................... | .................... |
| AA | 9060 hCoV-19/Denmark/DCGC-21842/2020\|EPI_ISL_758398\|202 | .................... | .................... | ..................... | .................... | .................... | .................... |
| AA | 9061 hCoV-19/Denmark/DCGC-21843/2020\|EPI_ISL_758399\|202 | .................... | .................... | ..T.................. | .................... | .................... | .................... |
| AA | 9062 hCoV-19/Denmark/DCGC-21327/2020\|EPI_ISL_759680\|202 | .................... | .................... | ..................... | .................... | .................... | .................... |
| AA | 9063 hCoV-19/Denmark/DCGC-21328/2020\|EPI_ISL_759681\|202 | .................... | .................... | ..................... | .................... | .................... | .................... |
| AA | 9064 hCoV-19/Denmark/DCGC-21329/2020\|EPI_ISL_759682\|202 | .................... | .................... | ..................... | .................... | .................... | .................... |
| AA | 9065 hCoV-19/Denmark/DCGC-21331/2020\|EPI_ISL_759683\|202 | .................... | .................... | ..................... | .................... | .................... | .................... |
| AA | 9066 hCoV-19/Denmark/DCGC-21332/2020\|EPI_ISL_759684\|202 | .................... | .................... | ..................... | .................... | .................... | .................... |
| AA | 9067 hCoV-19/Denmark/DCGC-21333/2020\|EPI_ISL_759685\|202 | .................... | .................... | ..................... | .................... | .................... | .................... |
| AA | 9068 hCoV-19/Denmark/DCGC-21334/2020\|EPI_ISL_759686\|202 | .................... | .................... | ..................... | .................... | .................... | .................... |
| AA | 9069 hCoV-19/Denmark/DCGC-21337/2020\|EPI_ISL_759687\|202 | .................... | .................... | ..................... | .................... | .................... | .................... |

| | | | | | | | | |
|---|---|---|---|---|---|---|---|---|
| AA | 9070 | hCoV-19/Denmark/DCGC-21338/2020\|EPI_ISL_759688\|202 | ···················· | ···················· | ···················· | ···················· | ···················· | ···················· |
| AA | 9071 | hCoV-19/Denmark/DCGC-21339/2020\|EPI_ISL_759689\|202 | ···················· | ···················· | ···················· | ···················· | ···················· | ···················· |
| AA | 9072 | hCoV-19/Denmark/DCGC-21341/2020\|EPI_ISL_759690\|202 | ···················· | ···················· | ···················· | ···················· | ···················· | ···················· |
| AA | 9073 | hCoV-19/Denmark/DCGC-21343/2020\|EPI_ISL_759691\|202 | ···················· | ···················· | ···················· | ···················· | ···················· | ···················· |
| AA | 9074 | hCoV-19/Denmark/DCGC-21345/2020\|EPI_ISL_759692\|202 | ···················· | ···················· | ···················· | ···················· | ···················· | ···················· |
| AA | 9075 | hCoV-19/Denmark/DCGC-21349/2020\|EPI_ISL_759693\|202 | ···················· | ···················· | ···················· | ···················· | ···················· | ···················· |
| AA | 9076 | hCoV-19/Denmark/DCGC-21351/2020\|EPI_ISL_759694\|202 | ···················· | ···················· | ···················· | ···················· | ···················· | ···················· |
| AA | 9077 | hCoV-19/Denmark/DCGC-21352/2020\|EPI_ISL_759695\|202 | ···················· | ···················· | ···················· | ···················· | ···················· | ···················· |
| AA | 9078 | hCoV-19/Denmark/DCGC-21353/2020\|EPI_ISL_759696\|202 | ···················· | ···················· | ···················· | ···················· | ···················· | ···················· |
| AA | 9079 | hCoV-19/Denmark/DCGC-21354/2020\|EPI_ISL_759697\|202 | ···················· | ···················· | ···················· | ···················· | ···················· | ···················· |
| AA | 9080 | hCoV-19/Denmark/DCGC-21356/2020\|EPI_ISL_759698\|202 | ···················· | ···················· | ···················· | ···················· | ···················· | ···················· |
| AA | 9081 | hCoV-19/Denmark/DCGC-21357/2020\|EPI_ISL_759699\|202 | ···················· | ·A·················· | ···················· | ···················· | ···················· | ···················· |
| AB | 1 | hCoV-19/England/ALDP-CB4A6B/2020\|EPI_ISL_761115\|20 | ···················· | ···················· | ···················· | ···················· | ···················· | ···················· |
| AB | 2 | hCoV-19/England/QEUH-CAD7D7/2020\|EPI_ISL_761116\|20 | ···················· | ···················· | ···················· | ···················· | ···················· | ···················· |
| AB | 3 | hCoV-19/England/ALDP-CB4B1D/2020\|EPI_ISL_761117\|20 | ···················· | ···················· | ···················· | ···················· | ···················· | ···················· |
| AB | 4 | hCoV-19/England/QEUH-CAD7E6/2020\|EPI_ISL_761118\|20 | ···················· | ···················· | ···················· | ···················· | ···················· | ···················· |
| AB | 5 | hCoV-19/England/ALDP-CB5E31/2020\|EPI_ISL_761119\|20 | ···················· | ···················· | ···················· | ···················· | ···················· | ···················· |
| AB | 6 | hCoV-19/England/ALDP-CB44D6/2020\|EPI_ISL_761124\|20 | ···················· | ···················· | ···················· | ···················· | ···················· | ···················· |
| AB | 7 | hCoV-19/England/ALDP-CB6841/2020\|EPI_ISL_761126\|20 | ···················· | ···················· | ···················· | ···················· | ···················· | ···················· |
| AB | 8 | hCoV-19/England/ALDP-CB59B8/2020\|EPI_ISL_761129\|20 | ···················· | ···················· | ···················· | ···················· | ···················· | ···················· |
| AB | 9 | hCoV-19/England/QEUH-CADD8A/2020\|EPI_ISL_761131\|20 | ···················· | ···················· | ···················· | ···················· | ······T············· | ···················· |
| AB | 10 | hCoV-19/England/ALDP-CB37B0/2020\|EPI_ISL_761137\|20 | ···················· | ···················· | ···················· | ···················· | ···················· | ···················· |
| AB | 11 | hCoV-19/England/ALDP-CB3ED9/2020\|EPI_ISL_761138\|20 | ···················· | ···················· | ···················· | ···················· | ···················· | ···················· |
| AB | 12 | hCoV-19/England/ALDP-CB6780/2020\|EPI_ISL_761140\|20 | ···················· | ···················· | ···················· | ···················· | ···················· | ···················· |
| AB | 13 | hCoV-19/England/ALDP-CB5D07/2020\|EPI_ISL_761141\|20 | ···················· | ·A·················· | ···················· | ···················· | ···················· | ···A················ |
| AB | 14 | hCoV-19/England/ALDP-CB3E9D/2020\|EPI_ISL_761142\|20 | ···················· | ···················· | ···················· | ···················· | ···················· | ···················· |
| AB | 15 | hCoV-19/England/ALDP-CB3EE8/2020\|EPI_ISL_761144\|20 | ···················· | ···················· | ···················· | ···················· | ······T············· | ···················· |
| AB | 16 | hCoV-19/England/ALDP-CB4D71/2020\|EPI_ISL_761146\|20 | ···················· | ···················· | ···················· | ···················· | ···················· | ···················· |
| AB | 17 | hCoV-19/England/ALDP-CB361D/2020\|EPI_ISL_761147\|20 | ···················· | ···················· | ···················· | ···················· | ···················· | ···················· |
| AB | 18 | hCoV-19/England/ALDP-CB34F5/2020\|EPI_ISL_761148\|20 | ···················· | ···················· | ···················· | ···················· | ···················· | ···················· |
| AB | 19 | hCoV-19/England/ALDP-CB3FF4/2020\|EPI_ISL_761150\|20 | ···················· | ···················· | ···················· | ···················· | ···················· | ···················· |
| AB | 20 | hCoV-19/England/ALDP-CB4078/2020\|EPI_ISL_761153\|20 | ···················· | ···················· | ···················· | ···················· | ···················· | ···················· |
| AB | 21 | hCoV-19/England/ALDP-CB35A7/2020\|EPI_ISL_761156\|20 | ···················· | ···················· | ···················· | ···················· | ···················· | ···················· |
| AB | 22 | hCoV-19/England/ALDP-CB3695/2020\|EPI_ISL_761163\|20 | ···················· | ···················· | ···················· | ···················· | ···················· | ···················· |
| AB | 23 | hCoV-19/England/ALDP-CB4685/2020\|EPI_ISL_761164\|20 | ···················· | ···················· | ···················· | ···················· | ···················· | ···················· |
| AB | 24 | hCoV-19/England/ALDP-CB352F/2020\|EPI_ISL_761166\|20 | ···················· | ···················· | ···················· | ···················· | ···················· | ···················· |
| AB | 25 | hCoV-19/England/ALDP-CB405A/2020\|EPI_ISL_761167\|20 | ···················· | ···················· | ···················· | ···················· | ······T············· | ···················· |
| AB | 26 | hCoV-19/England/ALDP-CB49E6/2020\|EPI_ISL_761169\|20 | ···················· | ···················· | ···················· | ···················· | ···················· | ···················· |
| AB | 27 | hCoV-19/England/ALDP-CB4D53/2020\|EPI_ISL_761170\|20 | ···················· | ···················· | ···················· | ···················· | ···················· | ···················· |
| AB | 28 | hCoV-19/England/ALDP-CB3C48/2020\|EPI_ISL_761174\|20 | ···················· | ···················· | ···················· | ···················· | ···················· | ···················· |
| AB | 29 | hCoV-19/England/ALDP-CB6762/2020\|EPI_ISL_761175\|20 | ···················· | ···················· | ···················· | ···················· | ···················· | ···················· |
| AB | 30 | hCoV-19/England/QEUH-CAD977/2020\|EPI_ISL_761176\|20 | ···················· | ···················· | ···················· | ···················· | ···········C········ | ···················· |
| AB | 31 | hCoV-19/England/ALDP-CB3F03/2020\|EPI_ISL_761178\|20 | ···················· | ···················· | ···················· | ···················· | ···················· | ···················· |
| AB | 32 | hCoV-19/England/ALDP-CB51FC/2020\|EPI_ISL_761179\|20 | ···················· | ···················· | ···················· | ···················· | ···················· | ···················· |
| AB | 33 | hCoV-19/England/ALDP-CB4D08/2020\|EPI_ISL_761180\|20 | ···················· | ···················· | ···················· | ···················· | ···················· | ···················· |
| AB | 34 | hCoV-19/England/ALDP-CB4CDE/2020\|EPI_ISL_761181\|20 | ···················· | ···················· | ···················· | ···················· | ···················· | ···················· |
| AB | 35 | hCoV-19/England/ALDP-CB36D1/2020\|EPI_ISL_761182\|20 | ···················· | ···················· | ···················· | ···················· | ···················· | ···················· |
| AB | 36 | hCoV-19/England/ALDP-CB4A89/2020\|EPI_ISL_761183\|20 | ···················· | ···················· | ···················· | ···················· | ···················· | ···················· |
| AB | 37 | hCoV-19/England/ALDP-CB48DA/2020\|EPI_ISL_761184\|20 | ···················· | ···················· | ···················· | ···················· | ···················· | ···················· |
| AB | 38 | hCoV-19/England/QEUH-CADE2D/2020\|EPI_ISL_761187\|20 | ···················· | ···················· | ···················· | ···················· | ···················· | ···················· |
| AB | 39 | hCoV-19/England/ALDP-CB40D2/2020\|EPI_ISL_761188\|20 | ···················· | ···················· | ···················· | ···················· | ···················· | ···················· |
| AB | 40 | hCoV-19/England/ALDP-CB4CFC/2020\|EPI_ISL_761189\|20 | ···················· | ···················· | ···················· | ···················· | ···················· | ···················· |
| AB | 41 | hCoV-19/England/ALDP-CB39C9/2020\|EPI_ISL_761190\|20 | ···················· | ···················· | ···················· | ···················· | ···················· | ···················· |
| AB | 42 | hCoV-19/England/QEUH-CADDD5/2020\|EPI_ISL_761191\|20 | ···················· | ···················· | ···················· | ···················· | ···················· | ···················· |
| AB | 43 | hCoV-19/England/ALDP-CB4AD4/2020\|EPI_ISL_761192\|20 | ···················· | ···················· | ···················· | ···················· | ···················· | ···················· |
| AB | 44 | hCoV-19/England/QEUH-CAD801/2020\|EPI_ISL_761194\|20 | ···················· | ···················· | ···················· | ···················· | ···················· | ···················· |
| AB | 45 | hCoV-19/England/ALDP-CB404B/2020\|EPI_ISL_761195\|20 | ···················· | ···················· | ···················· | ···················· | ···················· | ···················· |
| AB | 46 | hCoV-19/England/ALDP-CB6753/2020\|EPI_ISL_761196\|20 | ···················· | ···················· | ···················· | ···················· | ···················· | ···A················ |
| AB | 47 | hCoV-19/England/ALDP-CB6850/2020\|EPI_ISL_761199\|20 | ···················· | ···················· | ···················· | ···················· | ···················· | ···················· |
| AB | 48 | hCoV-19/England/MILK-CA4758/2020\|EPI_ISL_762445\|20 | ···················· | ···················· | ···················· | ···················· | ···················· | ···················· |
| AB | 49 | hCoV-19/England/MILK-CA9A0F/2020\|EPI_ISL_762450\|20 | ···················· | ···················· | ···················· | ···················· | ···················· | ···················· |
| AB | 50 | hCoV-19/England/MILK-CA3E72/2020\|EPI_ISL_762452\|20 | ···················· | ···················· | ···················· | ···················· | ···················· | ···················· |
| AB | 51 | hCoV-19/England/MILK-CA97BD/2020\|EPI_ISL_762464\|20 | ···················· | ···················· | ···················· | ···················· | ···················· | ···················· |
| AB | 52 | hCoV-19/England/MILK-CA4503/2020\|EPI_ISL_762468\|20 | ···T················ | ···················· | ···················· | ···················· | ···················· | ···················· |
| AB | 53 | hCoV-19/England/MILK-CA57EE/2020\|EPI_ISL_762469\|20 | ···················· | ···················· | ···················· | ···················· | ···················· | ···················· |
| AB | 54 | hCoV-19/England/MILK-CA4CD1/2020\|EPI_ISL_762482\|20 | ···················· | ···················· | ···················· | ···················· | ···················· | ···················· |
| AB | 55 | hCoV-19/England/MILK-CA4BE3/2020\|EPI_ISL_762486\|20 | ···················· | ···················· | ···················· | ···················· | ···················· | ···················· |
| AB | 56 | hCoV-19/England/MILK-CA9BC0/2020\|EPI_ISL_762487\|20 | ···················· | ···················· | ···················· | ···················· | ···················· | ···················· |
| AB | 57 | hCoV-19/England/MILK-CA9E9A/2020\|EPI_ISL_762489\|20 | ···················· | ···················· | ···················· | ···················· | ······T············· | ···················· |
| AB | 58 | hCoV-19/England/MILK-CA511D/2020\|EPI_ISL_762491\|20 | ···················· | ···················· | ···················· | ···················· | ···················· | ···················· |
| AB | 59 | hCoV-19/USA/CA-LACPHL-AF00063/2020\|EPI_ISL_766000\| | ···················· | ···················· | ···················· | ···················· | ···················· | ···················· |
| AB | 60 | hCoV-19/USA/CA-LACPHL-AF00064/2020\|EPI_ISL_766001\| | ···················· | ···················· | ···················· | ···················· | ···················· | ···················· |
| AB | 61 | hCoV-19/USA/CA-LACPHL-AF00065/2020\|EPI_ISL_766002\| | ···················· | ···················· | ···················· | ···················· | ···················· | ···················· |
| AB | 62 | hCoV-19/USA/CA-LACPHL-AF00066/2020\|EPI_ISL_766003\| | ···················· | ···················· | ···················· | ···················· | ···················· | ···················· |
| AB | 63 | hCoV-19/USA/CA-LACPHL-AF00067/2020\|EPI_ISL_766004\| | ···················· | ·A·················· | ···················· | ···················· | ···················· | ···················· |
| AB | 64 | hCoV-19/USA/CA-LACPHL-AF00068/2020\|EPI_ISL_766005\| | ···················· | ···················· | ···················· | ···················· | ···················· | ···················· |
| AB | 65 | hCoV-19/USA/CA-LACPHL-AF00069/2020\|EPI_ISL_766006\| | ···················· | ·A·················· | ···················· | ···················· | ···················· | ···················· |
| AB | 66 | hCoV-19/USA/CA-LACPHL-AF00070/2020\|EPI_ISL_766007\| | ···················· | ···················· | ···················· | ···················· | ···················· | ···················· |
| AB | 67 | hCoV-19/Wales/ALDP-C04BB4/2020\|EPI_ISL_767301\|2020 | ···················· | ···················· | ···················· | ···················· | ···················· | ···················· |
| AB | 68 | hCoV-19/Wales/ALDP-C04E42/2020\|EPI_ISL_767302\|2020 | ···················· | ···················· | ···················· | ···················· | ···················· | ···················· |

| | | | | | | | |
|---|---|---|---|---|---|---|---|
| AB | 69 hCoV-19/Wales/ALDP-C04DFA/2020\|EPI_ISL_767303\|2020 | ···················· | ···················· | ···················· | ···················· | ···················· | ···················· |
| AB | 70 hCoV-19/Wales/ALDP-C04BF0/2020\|EPI_ISL_767305\|2020 | ···················· | ···················· | ···················· | ···················· | ···················· | ···················· |
| AB | 71 hCoV-19/Wales/ALDP-C04DEB/2020\|EPI_ISL_767306\|2020 | ···················· | ···················· | ···················· | ···················· | ···················· | ···················· |
| AB | 72 hCoV-19/Wales/ALDP-C04D90/2020\|EPI_ISL_767307\|2020 | ···················· | ···················· | ···················· | ···················· | ···················· | ···················· |
| AB | 73 hCoV-19/Wales/ALDP-C04ECA/2020\|EPI_ISL_767308\|2020 | ···················· | ···················· | ···················· | ···················· | ···················· | ···················· |
| AB | 74 hCoV-19/Wales/ALDP-C04EF7/2020\|EPI_ISL_767309\|2020 | ···················· | ···················· | ···················· | ···················· | ···················· | ···················· |
| AB | 75 hCoV-19/Wales/ALDP-C04950/2020\|EPI_ISL_767311\|2020 | ···················· | ···················· | ···················· | ···················· | ···················· | ···················· |
| AB | 76 hCoV-19/Wales/ALDP-C04923/2020\|EPI_ISL_767312\|2020 | ···················· | ···················· | ···················· | ···················· | ···················· | ···················· |
| AB | 77 hCoV-19/Wales/ALDP-C04E15/2020\|EPI_ISL_767313\|2020 | ···················· | ···················· | ···················· | ···················· | ···················· | ···················· |
| AB | 78 hCoV-19/Wales/ALDP-C048F9/2020\|EPI_ISL_767314\|2020 | ···················· | ···················· | ···················· | ···················· | ···················· | ···················· |
| AB | 79 hCoV-19/Wales/ALDP-C0496F/2020\|EPI_ISL_767315\|2020 | ···················· | ···················· | ···················· | ···················· | ···················· | ···················· |
| AB | 80 hCoV-19/Wales/ALDP-C04B3C/2020\|EPI_ISL_767316\|2020 | ···················· | ···················· | ···················· | ···················· | ···················· | ···················· |
| AB | 81 hCoV-19/Wales/ALDP-C04932/2020\|EPI_ISL_767317\|2020 | ···················· | ···················· | ···················· | ···················· | ···················· | ···················· |
| AB | 82 hCoV-19/Wales/ALDP-C04905/2020\|EPI_ISL_767318\|2020 | ···················· | ···················· | ···················· | ···················· | ···················· | ···················· |
| AB | 83 hCoV-19/Wales/ALDP-BC8898/2020\|EPI_ISL_767319\|2020 | ···················· | ···················· | ···················· | ···················· | ···················· | ···················· |
| AB | 84 hCoV-19/Wales/ALDP-BC7680/2020\|EPI_ISL_767320\|2020 | ···················· | ···················· | ···················· | ···················· | ···················· | ···················· |
| AB | 85 hCoV-19/Wales/ALDP-BC7644/2020\|EPI_ISL_767321\|2020 | ···················· | ···················· | ···················· | ···················· | ···················· | ···················· |
| AB | 86 hCoV-19/Wales/ALDP-BC640F/2020\|EPI_ISL_767322\|2020 | ···················· | ···················· | ···················· | ···················· | ···················· | ···················· |
| AB | 87 hCoV-19/Wales/ALDP-BC769F/2020\|EPI_ISL_767323\|2020 | ···················· | ···················· | ···················· | ···················· | ···················· | ···················· |
| AB | 88 hCoV-19/Wales/ALDP-BC64B4/2020\|EPI_ISL_767324\|2020 | ···················· | ···················· | ···················· | ···················· | ····T··············· | ···················· |
| AB | 89 hCoV-19/Wales/ALDP-BC7671/2020\|EPI_ISL_767325\|2020 | ···················· | ···················· | ···················· | ···················· | ···················· | ···················· |
| AB | 90 hCoV-19/Wales/ALDP-BC641E/2020\|EPI_ISL_767326\|2020 | ···················· | ···················· | ···················· | ···················· | ···················· | ···················· |
| AB | 91 hCoV-19/Wales/ALDP-BC887A/2020\|EPI_ISL_767327\|2020 | ···················· | ···················· | ···················· | ···················· | ···················· | ···················· |
| AB | 92 hCoV-19/Wales/ALDP-BC88C5/2020\|EPI_ISL_767328\|2020 | ···················· | ···················· | ···················· | ···················· | ···················· | ···················· |
| AB | 93 hCoV-19/Wales/ALDP-BC6496/2020\|EPI_ISL_767329\|2020 | ···················· | ···················· | ···················· | ···················· | ···················· | ···················· |
| AB | 94 hCoV-19/Wales/ALDP-BC7653/2020\|EPI_ISL_767330\|2020 | ···················· | ···················· | ···················· | ···················· | ···················· | ···················· |
| AB | 95 hCoV-19/Wales/ALDP-BC88A7/2020\|EPI_ISL_767332\|2020 | ···················· | ···················· | ···················· | ···················· | ···················· | ···················· |
| AB | 96 hCoV-19/Wales/ALDP-BC63E4/2020\|EPI_ISL_767333\|2020 | ···················· | ···················· | ···················· | ···················· | ···················· | ···················· |
| AB | 97 hCoV-19/Australia/NSW1375/2020\|EPI_ISL_768600\|2020 | ···················· | ···················· | ···················· | ···················· | ···················· | ···················· |
| AB | 98 hCoV-19/Australia/NSW1377/2020\|EPI_ISL_768601\|2020 | ···················· | ···················· | ···················· | ···················· | ···················· | ···················· |
| AB | 99 hCoV-19/Singapore/1511/2020\|EPI_ISL_768627\|2020-12 | ···················· | ···················· | ···················· | ···················· | ···················· | ···················· |
| AB | 100 hCoV-19/Singapore/1512/2020\|EPI_ISL_768628\|2020-12 | ···················· | ···················· | ···················· | ···················· | ···················· | ···················· |
| AB | 101 hCoV-19/Singapore/1513/2020\|EPI_ISL_768629\|2020-12 | ···················· | ···················· | ···················· | ···················· | ···················· | ···················· |
| AB | 102 hCoV-19/Singapore/1514/2020\|EPI_ISL_768630\|2020-12 | ···················· | ···················· | ···················· | ···················· | ···················· | ···················· |
| AB | 103 hCoV-19/Germany/TH-IIMK-CaSe-29/2020\|EPI_ISL_76863 | ···················· | ···················· | ···················· | ···················· | ···················· | ···················· |
| AB | 104 hCoV-19/Japan/IC-0425/2020\|EPI_ISL_768634\|2020-11 | ···················· | ···················· | ···················· | ···················· | ···················· | ···················· |
| AB | 105 hCoV-19/Japan/IC-0426/2020\|EPI_ISL_768635\|2020-11 | ···················· | ···················· | ···················· | ···················· | ···················· | ···················· |
| AB | 106 hCoV-19/Japan/IC-0427/2020\|EPI_ISL_768636\|2020-11 | ···················· | ···················· | ···················· | ···················· | ···················· | ···················· |
| AB | 107 hCoV-19/Japan/IC-0428/2020\|EPI_ISL_768637\|2020-11 | ···················· | ···················· | ···················· | ···················· | ···················· | ···················· |
| AB | 108 hCoV-19/Japan/IC-0429/2020\|EPI_ISL_768638\|2020-11 | ···················· | ···················· | ···················· | ···················· | ···················· | ···················· |
| AB | 109 hCoV-19/Japan/IC-0430/2020\|EPI_ISL_768639\|2020-11 | ···················· | ···················· | ···················· | ···················· | ···················· | ···················· |
| AB | 110 hCoV-19/Japan/IC-0431/2020\|EPI_ISL_768640\|2020-12 | ···················· | ···················· | ···················· | ···················· | ···················· | ···················· |
| AB | 111 hCoV-19/Japan/IC-0432/2020\|EPI_ISL_768641\|2020-12 | ···················· | ···················· | ···················· | ···················· | ···················· | ···················· |
| AB | 112 hCoV-19/Japan/IC-0434/2020\|EPI_ISL_768643\|2020-09 | ···················· | ···················· | ···················· | ···················· | ···················· | ···················· |
| AB | 113 hCoV-19/Japan/IC-0435/2020\|EPI_ISL_768644\|2020-09 | ···················· | ···················· | ···················· | ···················· | ···················· | ···················· |
| AB | 114 hCoV-19/Japan/IC-0436/2020\|EPI_ISL_768645\|2020-09 | ···················· | ···················· | ···················· | ···················· | ···················· | ···················· |
| AB | 115 hCoV-19/Japan/IC-0437/2020\|EPI_ISL_768646\|2020-10 | ···················· | ···················· | ···················· | ···················· | ···················· | ···················· |
| AB | 116 hCoV-19/Japan/IC-0438/2020\|EPI_ISL_768647\|2020-10 | ···················· | ···················· | ···················· | ···················· | ···················· | ···················· |
| AB | 117 hCoV-19/Japan/IC-0439/2020\|EPI_ISL_768648\|2020-10 | ···················· | ···················· | ···················· | ···················· | ···················· | ···················· |
| AB | 118 hCoV-19/Japan/IC-0440/2020\|EPI_ISL_768649\|2020-10 | ···················· | ···················· | ···················· | ···················· | ···················· | ···················· |
| AB | 119 hCoV-19/Japan/IC-0441/2020\|EPI_ISL_768650\|2020-10 | ···················· | ·A·················· | ···················· | ···················· | ···················· | ···················· |
| AB | 120 hCoV-19/Japan/IC-0442/2020\|EPI_ISL_768651\|2020-11 | ···················· | ···················· | ···················· | ···················· | ···················· | ···················· |
| AB | 121 hCoV-19/Japan/IC-0443/2020\|EPI_ISL_768652\|2020-11 | ···················· | ···················· | ···················· | ···················· | ···················· | ···················· |
| AB | 122 hCoV-19/Japan/IC-0444/2020\|EPI_ISL_768653\|2020-11 | ···················· | ···················· | ···················· | ···················· | ···················· | ···················· |
| AB | 123 hCoV-19/Japan/IC-0445/2020\|EPI_ISL_768654\|2020-11 | ···················· | ···················· | ···················· | ···················· | ···················· | ···················· |
| AB | 124 hCoV-19/Japan/IC-0447/2020\|EPI_ISL_768656\|2020-12 | ···················· | ···················· | ···················· | ···················· | ···················· | ···················· |
| AB | 125 hCoV-19/Japan/IC-0448/2020\|EPI_ISL_768657\|2020-12 | ···················· | ···················· | ···················· | ···················· | ···················· | ···················· |
| AB | 126 hCoV-19/Japan/IC-0450/2020\|EPI_ISL_768659\|2020-10 | ···················· | ···················· | ···················· | ···················· | ···················· | ···················· |
| AB | 127 hCoV-19/Japan/IC-0451/2020\|EPI_ISL_768660\|2020-10 | ···················· | ···················· | ···················· | ···················· | ···················· | ···················· |
| AB | 128 hCoV-19/Japan/IC-0452/2020\|EPI_ISL_768661\|2020-12 | ···················· | ··········A········· | ···················· | ···················· | ···················· | ···················· |
| AB | 129 hCoV-19/Japan/IC-0453/2020\|EPI_ISL_768662\|2020-12 | ···················· | ···················· | ···················· | ···················· | ···················· | ···················· |
| AB | 130 hCoV-19/Japan/IC-0457/2020\|EPI_ISL_768666\|2020-12 | ···················· | ···················· | ···················· | ···················· | ···················· | ···················· |
| AB | 131 hCoV-19/Japan/IC-0458/2020\|EPI_ISL_768667\|2020-09 | ···················· | ···················· | ···················· | ···················· | ···················· | ···················· |
| AB | 132 hCoV-19/Japan/IC-0459/2020\|EPI_ISL_768668\|2020-12 | ···················· | ·A·················· | ···················· | ···················· | ···················· | ···A················ |
| AB | 133 hCoV-19/Japan/IC-0460/2020\|EPI_ISL_768669\|2020-12 | ···················· | ····A··············· | ···················· | ···················· | ···················· | ···················· |
| AB | 134 hCoV-19/USA/WY-WPHL-20061934/2020\|EPI_ISL_769900\|2 | ···················· | ···················· | ···················· | ···················· | ···················· | ···················· |
| AB | 135 hCoV-19/USA/WY-WPHL-20061902/2020\|EPI_ISL_769902\|2 | ···················· | ···················· | ···················· | ···················· | ···················· | ···················· |
| AB | 136 hCoV-19/USA/WY-WPHL-20061888/2020\|EPI_ISL_769903\|2 | ···················· | ···················· | ···················· | ···················· | ···················· | ···················· |
| AB | 137 hCoV-19/USA/WY-WPHL-20061820/2020\|EPI_ISL_769904\|2 | ···················· | ··C················· | ···················· | ···················· | ···················· | ···················· |
| AB | 138 hCoV-19/USA/WY-WPHL-20061819/2020\|EPI_ISL_769905\|2 | ···················· | ·A·················· | ···················· | ···················· | ···················· | ···················· |
| AB | 139 hCoV-19/USA/WY-WPHL-20044834/2020\|EPI_ISL_769907\|2 | ···················· | ···················· | ···················· | ···················· | ···················· | ···················· |
| AB | 140 hCoV-19/USA/WY-WPHL-20044813/2020\|EPI_ISL_769908\|2 | ···················· | ···················· | ···················· | ···················· | ···················· | ···················· |
| AB | 141 hCoV-19/USA/WY-WPHL-20044665/2020\|EPI_ISL_769909\|2 | ···················· | ···················· | ···················· | ···················· | ···················· | ···················· |
| AB | 142 hCoV-19/USA/WY-WPHL-20044583/2020\|EPI_ISL_769910\|2 | ···················· | ···················· | ···················· | ···················· | ···················· | ···················· |
| AB | 143 hCoV-19/USA/WY-WPHL-20044559/2020\|EPI_ISL_769911\|2 | ···················· | ···················· | ···················· | ···················· | ···················· | ···················· |
| AB | 144 hCoV-19/USA/WY-WPHL-20044555/2020\|EPI_ISL_769912\|2 | ···················· | ·A·················· | ···················· | ···················· | ···················· | ···················· |
| AB | 145 hCoV-19/USA/WY-WPHL-20044542/2020\|EPI_ISL_769913\|2 | ···················· | ···················· | ···················· | ···················· | ···················· | ···················· |
| AB | 146 hCoV-19/USA/WY-WPHL-20044526/2020\|EPI_ISL_769914\|2 | ···················· | ···················· | ···················· | ···················· | ···················· | ···················· |
| AB | 147 hCoV-19/USA/WY-WPHL-20044519/2020\|EPI_ISL_769915\|2 | ···················· | ···················· | ···················· | ···················· | ···················· | ···················· |
| AB | 148 hCoV-19/USA/NY-Wadsworth-292763-01/2020\|EPI_ISL_76 | ···················· | ···················· | ···················· | ···················· | ···················· | ···················· |

| | | | | | | | | |
|---|---|---|---|---|---|---|---|---|
| AB | 149 hCoV-19/USA/NY-Wadsworth-292759-01/2020\|EPI_ISL_76 | ···················· | ·A·················· | ···················· | ···················· | ···················· | ···················· | |
| AB | 150 hCoV-19/USA/NY-Wadsworth-292810-01/2020\|EPI_ISL_76 | ···················· | ···················· | ···················· | ···················· | ···················· | ···················· | |
| AB | 151 hCoV-19/USA/NY-Wadsworth-292738-01/2020\|EPI_ISL_76 | ···················· | ·A·················· | ···················· | ···················· | ···················· | ···················· | |
| AB | 152 hCoV-19/USA/NY-Wadsworth-292744-01/2020\|EPI_ISL_76 | ···················· | ···················· | ···················· | ···················· | ···················· | ···················· | |
| AB | 153 hCoV-19/USA/NY-Wadsworth-292745-01/2020\|EPI_ISL_76 | ···················· | ···················· | ···················· | ···················· | ···················· | ···················· | |
| AB | 154 hCoV-19/USA/NY-Wadsworth-292752-01/2020\|EPI_ISL_76 | ···················· | ···················· | ···················· | ···················· | ···················· | ···················· | |
| AB | 155 hCoV-19/USA/NY-Wadsworth-292753-01/2020\|EPI_ISL_76 | ··················T· | ···················· | ···················· | ···················· | ···················· | ···················· | 1 |
| AB | 156 hCoV-19/USA/NY-Wadsworth-292754-01/2020\|EPI_ISL_76 | ···················· | ·A·················· | ···················· | ···················· | ···················· | ···················· | |
| AB | 157 hCoV-19/USA/NY-Wadsworth-292756-01/2020\|EPI_ISL_76 | ···················· | ···················· | ···················· | ···················· | ···················· | ···················· | |
| AB | 158 hCoV-19/USA/NY-Wadsworth-292757-01/2020\|EPI_ISL_76 | ···················· | ···················· | ···················· | ···················· | ···················· | ···················· | |
| AB | 159 hCoV-19/USA/NY-Wadsworth-292758-01/2020\|EPI_ISL_76 | ···················· | ···················· | ···················· | ···················· | ···················· | ···················· | |
| AB | 160 hCoV-19/USA/NY-Wadsworth-292792-01/2020\|EPI_ISL_76 | ···················· | ·A·················· | ···················· | ···················· | ···················· | ···················· | |
| AB | 161 hCoV-19/USA/NY-Wadsworth-292793-01/2020\|EPI_ISL_76 | ···················· | ·A·················· | ···················· | ···················· | ···················· | ···················· | |
| AB | 162 hCoV-19/USA/NY-Wadsworth-292794-01/2020\|EPI_ISL_76 | ···················· | ···················· | ···················· | ···················· | ···················· | ···················· | |
| AB | 163 hCoV-19/USA/NY-Wadsworth-292795-01/2020\|EPI_ISL_76 | ···················· | ···················· | ···················· | ···················· | ···················· | ···················· | |
| AB | 164 hCoV-19/USA/NY-Wadsworth-292797-01/2020\|EPI_ISL_76 | ···················· | ···················· | ···················· | ···················· | ···················· | ···················· | |
| AB | 165 hCoV-19/USA/NY-Wadsworth-292800-01/2020\|EPI_ISL_76 | ···················· | ·A·················· | ···················· | ···················· | ···················· | ···················· | |
| AB | 166 hCoV-19/USA/NY-Wadsworth-292803-01/2020\|EPI_ISL_76 | ···················· | ···················· | ···················· | ···················· | ···················· | ···················· | |
| AB | 167 hCoV-19/USA/NY-Wadsworth-292805-01/2020\|EPI_ISL_76 | ···················· | ·A·················· | ···················· | ···················· | ···················· | ···················· | |
| AB | 168 hCoV-19/USA/NY-Wadsworth-292806-01/2020\|EPI_ISL_76 | ···················· | ···················· | ···················· | ···················· | ···················· | ···················· | |
| AB | 169 hCoV-19/USA/NY-Wadsworth-292807-01/2020\|EPI_ISL_76 | ···················· | ·A·················· | ···················· | ···················· | ···················· | ···················· | |
| AB | 170 hCoV-19/USA/NY-Wadsworth-292811-01/2020\|EPI_ISL_76 | ···················· | ·A·················· | ···················· | ···················· | ···················· | ···················· | |
| AB | 171 hCoV-19/USA/NY-Wadsworth-292812-01/2020\|EPI_ISL_76 | ···················· | ·A·················· | ···················· | ···················· | ···················· | ···················· | |
| AB | 172 hCoV-19/USA/NY-Wadsworth-292814-01/2020\|EPI_ISL_76 | ···················· | ···················· | ···················· | ···················· | ···················· | ···················· | |
| AB | 173 hCoV-19/USA/NY-Wadsworth-292820-01/2020\|EPI_ISL_76 | ···················· | ···················· | ···················· | ···················· | ···················· | ···················· | |
| AB | 174 hCoV-19/USA/NY-Wadsworth-292823-01/2020\|EPI_ISL_76 | ···················· | ·R·················· | ···················· | ···················· | ···················· | ···················· | |
| AB | 175 hCoV-19/USA/NY-Wadsworth-292824-01/2020\|EPI_ISL_76 | ···················· | ···················· | ···················· | ···················· | ···················· | ···················· | |
| AB | 176 hCoV-19/USA/NY-Wadsworth-292826-01/2020\|EPI_ISL_76 | ···················· | ···················· | ···················· | ···················· | ···················· | ···················· | |
| AB | 177 hCoV-19/USA/NY-Wadsworth-292830-01/2020\|EPI_ISL_76 | ···················· | ·A·················· | ···················· | ···················· | ···················· | ···················· | |
| AB | 178 hCoV-19/USA/NY-Wadsworth-292832-01/2020\|EPI_ISL_76 | ···················· | ···················· | ···················· | ···················· | ···················· | ···················· | |
| AB | 179 hCoV-19/USA/NY-Wadsworth-292834-01/2020\|EPI_ISL_76 | ···················· | ···················· | ···················· | ···················· | ···················· | ···················· | |
| AB | 180 hCoV-19/USA/NY-Wadsworth-292839-01/2020\|EPI_ISL_76 | ···················· | ···················· | ···················· | ···················· | ···················· | ···················· | |
| AB | 181 hCoV-19/USA/NY-Wadsworth-292840-01/2020\|EPI_ISL_76 | ···················· | ·A·················· | ···················· | ···················· | ···················· | ···················· | |
| AB | 183 hCoV-19/USA/NY-Wadsworth-292843-01/2020\|EPI_ISL_76 | ···················· | ·A·················· | ···················· | ···················· | ···················· | ···················· | |
| AB | 184 hCoV-19/USA/NY-Wadsworth-292844-01/2020\|EPI_ISL_76 | ···················· | ···················· | ···················· | ···················· | ···················· | ···················· | |
| AB | 185 hCoV-19/USA/NY-Wadsworth-292845-01/2020\|EPI_ISL_76 | ···················· | ···················· | ···················· | ···················· | ···················· | ···················· | |
| AB | 186 hCoV-19/USA/NY-Wadsworth-292964-01/2020\|EPI_ISL_76 | ···················· | ···················· | ···················· | ···················· | ···················· | ···················· | |
| AB | 187 hCoV-19/USA/NY-Wadsworth-292966-01/2020\|EPI_ISL_76 | ···················· | ·A·················· | ···················· | ···················· | ···················· | ···················· | |
| AB | 188 hCoV-19/Costa Rica/INC-0162/2020\|EPI_ISL_769987\|20 | ···················· | ···················· | ··A················· | ···················· | ···················· | ···················· | |
| AB | 189 hCoV-19/Costa Rica/INC-0178/2020\|EPI_ISL_769988\|20 | ···················· | ···················· | ···················· | ···················· | ···················· | ···················· | |
| AB | 190 hCoV-19/Costa Rica/INC-0176/2020\|EPI_ISL_769989\|20 | ···················· | ···················· | ···················· | ···················· | ···················· | ··A················· | |
| AB | 191 hCoV-19/Costa Rica/INC-0144/2020\|EPI_ISL_769990\|20 | ···················· | ···················· | ···················· | ···················· | ···················· | ···················· | |
| AB | 192 hCoV-19/Costa Rica/INC-0148/2020\|EPI_ISL_769991\|20 | ···················· | ···················· | ···················· | ···················· | ···················· | ···················· | |
| AB | 193 hCoV-19/Costa Rica/INC-0155/2020\|EPI_ISL_769992\|20 | ···················· | ···················· | ···················· | ···················· | ········A··········· | ···················· | |
| AB | 194 hCoV-19/Costa Rica/INC-0143/2020\|EPI_ISL_769993\|20 | ···················· | ···················· | ···················· | ···················· | ···················· | ···················· | |
| AB | 195 hCoV-19/Costa Rica/INC-0146/2020\|EPI_ISL_769994\|20 | ···················· | ···················· | ···················· | ···················· | ···················· | ···················· | |
| AB | 196 hCoV-19/Costa Rica/INC-0154/2020\|EPI_ISL_769995\|20 | ···················· | ···················· | ···················· | ···················· | ···················· | ···················· | |
| AB | 197 hCoV-19/Costa Rica/INC-0163/2020\|EPI_ISL_769996\|20 | ···················· | ···················· | ···················· | ···················· | ···················· | ···················· | |
| AB | 198 hCoV-19/Costa Rica/INC-0172/2020\|EPI_ISL_769997\|20 | ···················· | ···················· | ···················· | ···················· | ···················· | ···················· | |
| AB | 199 hCoV-19/Costa Rica/INC-0174/2020\|EPI_ISL_769998\|20 | ···················· | ···················· | ···················· | ···················· | ···················· | ···················· | |
| AB | 200 hCoV-19/Costa Rica/INC-0186/2020\|EPI_ISL_769999\|20 | ···················· | ···················· | ···················· | ···················· | ···················· | ···················· | |
| AB | 201 hCoV-19/Luxembourg/LNS1337786/2020\|EPI_ISL_770900\| | ···················· | ···················· | ···················· | ···················· | ···················· | ···················· | |
| AB | 202 hCoV-19/Luxembourg/LNS8811428/2020\|EPI_ISL_770901\| | ···················· | ···················· | ···················· | ···················· | ···················· | ···················· | |
| AB | 203 hCoV-19/Luxembourg/LNS5447190/2020\|EPI_ISL_770902\| | ···················· | ···················· | ···················· | ···················· | ···················· | ···················· | |
| AB | 204 hCoV-19/Luxembourg/LNS6901259/2020\|EPI_ISL_770903\| | ···················· | ···················· | ···················· | ···················· | ···················· | ···················· | |
| AB | 205 hCoV-19/Luxembourg/LNS5759444/2020\|EPI_ISL_770904\| | ···················· | ···················· | ···················· | ···················· | ···················· | ···················· | |
| AB | 206 hCoV-19/Luxembourg/LNS1568478/2020\|EPI_ISL_770910\| | ···················· | ···················· | ···················· | ···················· | ···················· | ···················· | |
| AB | 207 hCoV-19/Luxembourg/LNS4839843/2020\|EPI_ISL_770911\| | ···················· | ···················· | ···················· | ···················· | ···················· | ···················· | |
| AB | 208 hCoV-19/Luxembourg/LNS6836311/2020\|EPI_ISL_770912\| | ···················· | ···A················ | ···················· | ···················· | ···················· | ···················· | |
| AB | 209 hCoV-19/Luxembourg/LNS9066840/2020\|EPI_ISL_770913\| | ···················· | ···················· | ···················· | ···················· | ···················· | ···················· | |
| AB | 210 hCoV-19/Luxembourg/LNS7652114/2020\|EPI_ISL_770914\| | ···················· | ···················· | ···················· | ···················· | ···················· | ···················· | |
| AB | 211 hCoV-19/Luxembourg/LNS5755279/2020\|EPI_ISL_770915\| | ···················· | ···················· | ···················· | ···················· | ···················· | ···················· | |
| AB | 212 hCoV-19/Luxembourg/LNS6971732/2020\|EPI_ISL_770916\| | ···················· | ···················· | ···················· | ···················· | ···················· | ···················· | |
| AB | 213 hCoV-19/Luxembourg/LNS8526074/2020\|EPI_ISL_770917\| | ···················· | ···················· | ···················· | ···················· | ···················· | ···················· | |
| AB | 214 hCoV-19/Luxembourg/LNS9352817/2020\|EPI_ISL_770918\| | ···················· | ···················· | ···················· | ···················· | ···················· | ···················· | |
| AB | 215 hCoV-19/Luxembourg/LNS6243765/2020\|EPI_ISL_770919\| | ···················· | ···················· | ···················· | ···················· | ···················· | ···················· | |
| AB | 216 hCoV-19/Luxembourg/LNS5239699/2020\|EPI_ISL_770940\| | ···················· | ···················· | ···················· | ···················· | ···················· | ···················· | |
| AB | 217 hCoV-19/Luxembourg/LNS0432074/2020\|EPI_ISL_770941\| | ···················· | ···················· | ···················· | ···················· | ···················· | ···················· | |
| AB | 218 hCoV-19/Luxembourg/LNS0655881/2020\|EPI_ISL_770942\| | ···················· | ···················· | ···················· | ···················· | ···················· | ···················· | |
| AB | 219 hCoV-19/Luxembourg/LNS6616225/2020\|EPI_ISL_770943\| | ···················· | ···················· | ···················· | ···················· | ···················· | ···················· | |
| AB | 220 hCoV-19/Luxembourg/LNS6035285/2020\|EPI_ISL_770944\| | ···················· | ···················· | ···················· | ···················· | ···················· | ···················· | |
| AB | 221 hCoV-19/Luxembourg/LNS6060441/2020\|EPI_ISL_770945\| | ···················· | ···················· | ···················· | ···················· | ···················· | ···················· | |
| AB | 222 hCoV-19/Luxembourg/LNS8805109/2020\|EPI_ISL_770946\| | ···················· | ···················· | ···················· | ···················· | ···················· | ···················· | |
| AB | 223 hCoV-19/Luxembourg/LNS2751620/2020\|EPI_ISL_770947\| | ···················· | ···················· | ···················· | ···················· | ···················· | ···················· | |
| AB | 224 hCoV-19/Luxembourg/LNS6504106/2020\|EPI_ISL_770948\| | ···················· | ···················· | ···················· | ···················· | ···················· | ···················· | |
| AB | 225 hCoV-19/Luxembourg/LNS5868607/2020\|EPI_ISL_770949\| | ···················· | ···················· | ···················· | ···················· | ···················· | ···················· | |
| AB | 226 hCoV-19/Luxembourg/LNS0891389/2020\|EPI_ISL_770950\| | ···················· | ···················· | ···················· | ···················· | ···················· | ···················· | |
| AB | 227 hCoV-19/Luxembourg/LNS5745708/2020\|EPI_ISL_770951\| | ···················· | ···················· | ···················· | ···················· | ···················· | ···················· | |
| AB | 228 hCoV-19/Luxembourg/LNS5821163/2020\|EPI_ISL_770952\| | ···················· | ···················· | ···················· | ···················· | ···················· | ···················· | |
| AB | 229 hCoV-19/Luxembourg/LNS1649454/2020\|EPI_ISL_770953\| | ···················· | ···················· | ···················· | ···················· | ···················· | ···················· | |

| | | | | | | | | | |
|---|---|---|---|---|---|---|---|---|---|
| AB | 230 | hCoV-19/Luxembourg/LNS0490955/2020\|EPI_ISL_770954\| | ···················· | ···················· | ····················· | ···················· | ···················· | ···················· | |
| AB | 231 | hCoV-19/Luxembourg/LNS8056927/2020\|EPI_ISL_770955\| | ···················· | ···················· | ····················· | ···················· | ···················· | ···················· | |
| AB | 232 | hCoV-19/Luxembourg/LNS4288043/2020\|EPI_ISL_770956\| | ···················· | ···················· | ····················· | ···················· | ···················· | ···················· | |
| AB | 233 | hCoV-19/Luxembourg/LNS3614652/2020\|EPI_ISL_770957\| | ···················· | ···················· | ····················· | ···················· | ···················· | ···················· | |
| AB | 234 | hCoV-19/Luxembourg/LNS9089301/2020\|EPI_ISL_770958\| | ···················· | ···················· | ····················· | ···················· | ···················· | ···················· | |
| AB | 235 | hCoV-19/Luxembourg/LNS7128997/2020\|EPI_ISL_770959\| | ···················· | ···················· | ····················· | ···················· | ···················· | ···················· | |
| AB | 236 | hCoV-19/Luxembourg/LNS6999817/2020\|EPI_ISL_770960\| | ···················· | ···················· | ····················· | ···················· | ···················· | ···················· | |
| AB | 237 | hCoV-19/Luxembourg/LNS5779400/2020\|EPI_ISL_770961\| | ···················· | ···················· | ····················· | ···················· | ···················· | ···················· | |
| AB | 238 | hCoV-19/Luxembourg/LNS7574226/2020\|EPI_ISL_770962\| | ···················· | ···················· | ····················· | ···················· | ···················· | ···················· | |
| AB | 239 | hCoV-19/Luxembourg/LNS3188060/2020\|EPI_ISL_770963\| | ···················· | ···················· | ····················· | ···················· | ···················· | ···················· | |
| AB | 240 | hCoV-19/Luxembourg/LNS3929243/2020\|EPI_ISL_770964\| | ···················· | ···················· | ····················· | ···················· | ···················· | ···················· | |
| AB | 241 | hCoV-19/Luxembourg/LNS2332603/2020\|EPI_ISL_770965\| | ···················· | ···················· | ····················· | ···················· | ···················· | ···················· | |
| AB | 242 | hCoV-19/Luxembourg/LNS6842253/2020\|EPI_ISL_770966\| | ···················· | ···················· | ····················· | ···················· | ···················· | ···················· | |
| AB | 243 | hCoV-19/Luxembourg/LNS1204905/2020\|EPI_ISL_770967\| | ···················· | ···················· | ····················· | ···················· | ···················· | ···················· | |
| AB | 244 | hCoV-19/Luxembourg/LNS0876307/2020\|EPI_ISL_770968\| | ···················· | ···················· | ····················· | ···················· | ···················· | ···················· | |
| AB | 245 | hCoV-19/Luxembourg/LNS7903820/2020\|EPI_ISL_770969\| | ···················· | ···················· | ··········T·········· | ···················· | ···················· | ···················· | |
| AB | 246 | hCoV-19/Luxembourg/LNS9306487/2020\|EPI_ISL_770970\| | ···················· | ···················· | ····················· | ···················· | ···················· | ···················· | |
| AB | 247 | hCoV-19/Luxembourg/LNS1573955/2020\|EPI_ISL_770971\| | ···················· | ···················· | ····················· | ···················· | ···················· | ···················· | |
| AB | 248 | hCoV-19/Luxembourg/LNS7078029/2020\|EPI_ISL_770972\| | ···················· | ···················· | ····················· | ···················· | ···················· | ···················· | |
| AB | 249 | hCoV-19/Luxembourg/LNS2686590/2020\|EPI_ISL_770973\| | ···················· | ···················· | ····················· | ···················· | ···················· | ···················· | |
| AB | 250 | hCoV-19/Luxembourg/LNS8740206/2020\|EPI_ISL_770974\| | ···················· | ···················· | ····················· | ···················· | ···················· | ···················· | |
| AB | 251 | hCoV-19/Luxembourg/LNS6167037/2020\|EPI_ISL_770975\| | ···················· | ···················· | ····················· | ···················· | ···················· | ···················· | |
| AB | 252 | hCoV-19/Luxembourg/LNS8554607/2020\|EPI_ISL_770976\| | ···················· | ···················· | ····················· | ···················· | ···················· | ···················· | |
| AB | 253 | hCoV-19/Luxembourg/LNS9787705/2020\|EPI_ISL_770977\| | ···················· | ····A··············· | ····················· | ···················· | ···················· | ···················· | |
| AB | 254 | hCoV-19/Luxembourg/LNS7619358/2020\|EPI_ISL_770978\| | ···················· | ···················· | ····················· | ···················· | ···················· | ···················· | |
| AB | 255 | hCoV-19/Luxembourg/LNS4296081/2020\|EPI_ISL_770979\| | ···················· | ···················· | ····················· | ···················· | ···················· | ···················· | |
| AB | 256 | hCoV-19/Luxembourg/LNS9749716/2020\|EPI_ISL_770980\| | ···················· | ···················· | ····················· | ···················· | ···················· | ···················· | |
| AB | 257 | hCoV-19/England/ALDP-CB3EF7/2020\|EPI_ISL_761200\|20 | ···················· | ···················· | ····················· | ···················· | ···················· | ···················· | |
| AB | 258 | hCoV-19/England/ALDP-CB3853/2020\|EPI_ISL_761201\|20 | ···················· | ···················· | ····················· | ···················· | ···················· | ···················· | |
| AB | 259 | hCoV-19/England/QEUH-CAD810/2020\|EPI_ISL_761204\|20 | ···················· | ···················· | ····················· | ···················· | ···················· | ···················· | |
| AB | 260 | hCoV-19/England/ALDP-CB40F0/2020\|EPI_ISL_761205\|20 | ···················· | ···················· | ····················· | ···················· | ···················· | ···················· | |
| AB | 261 | hCoV-19/England/ALDP-CB67EA/2020\|EPI_ISL_761206\|20 | ···················· | ···················· | ····················· | ···················· | ···················· | ···················· | |
| AB | 262 | hCoV-19/England/QEUH-CADCD8/2020\|EPI_ISL_761207\|20 | ··················T· | ···················· | ····················· | ···················· | ···················· | ···················· | 1 |
| AB | 263 | hCoV-19/Wales/ALDP-CB3BD2/2020\|EPI_ISL_761208\|2020 | ···················· | ···················· | ····················· | ···················· | ···················· | ···················· | |
| AB | 264 | hCoV-19/England/QEUH-CAD8F2/2020\|EPI_ISL_761209\|20 | ···················· | ···················· | ····················· | ···················· | ···················· | ···················· | |
| AB | 265 | hCoV-19/England/ALDP-CB3598/2020\|EPI_ISL_761210\|20 | ···················· | ·········T·········· | ····················· | ···················· | ···················· | ···················· | |
| AB | 266 | hCoV-19/Wales/ALDP-CB38CC/2020\|EPI_ISL_761211\|2020 | ···················· | ···················· | ····················· | ···················· | ···················· | ···················· | |
| AB | 267 | hCoV-19/England/ALDP-CB44C7/2020\|EPI_ISL_761212\|20 | ···················· | ···················· | ····················· | ···················· | ···················· | ···················· | |
| AB | 268 | hCoV-19/England/ALDP-CB688D/2020\|EPI_ISL_761215\|20 | ···················· | ·A·················· | ····················· | ···················· | ···················· | ···A················ | |
| AB | 269 | hCoV-19/England/ALDP-CB35C5/2020\|EPI_ISL_761216\|20 | ···················· | ···················· | ····················· | ···················· | ···················· | ···················· | |
| AB | 270 | hCoV-19/England/ALDP-CB6735/2020\|EPI_ISL_761219\|20 | ···················· | ·A·················· | ····················· | ···················· | ···················· | ···A················ | |
| AB | 271 | hCoV-19/England/ALDP-CB3862/2020\|EPI_ISL_761220\|20 | ···················· | ···················· | ····················· | ···················· | ···················· | ···················· | |
| AB | 272 | hCoV-19/England/ALDP-CB48AD/2020\|EPI_ISL_761222\|20 | ···················· | ···················· | ····················· | ···················· | ···················· | ···················· | |
| AB | 273 | hCoV-19/England/ALDP-CB4CED/2020\|EPI_ISL_761223\|20 | ···················· | ···················· | ····················· | ···················· | ···················· | ···················· | |
| AB | 274 | hCoV-19/England/QEUH-CADCAB/2020\|EPI_ISL_761225\|20 | ···················· | ···················· | ····················· | ···················· | ···················· | ···················· | |
| AB | 275 | hCoV-19/England/QEUH-CADCC9/2020\|EPI_ISL_761227\|20 | ···················· | ···················· | ····················· | ···················· | ···················· | ···················· | |
| AB | 276 | hCoV-19/England/ALDP-CB3808/2020\|EPI_ISL_761231\|20 | ···················· | ···················· | ····················· | ···················· | ···················· | ···················· | |
| AB | 277 | hCoV-19/England/ALDP-CB652C/2020\|EPI_ISL_761234\|20 | ···················· | ···················· | ····················· | ···················· | ···················· | ···················· | |
| AB | 278 | hCoV-19/England/ALDP-CB3835/2020\|EPI_ISL_761235\|20 | ···················· | ···················· | ··········T·········· | ···················· | ······T············· | ···················· | |
| AB | 279 | hCoV-19/England/ALDP-CB354D/2020\|EPI_ISL_761238\|20 | ···················· | ···················· | ····················· | ···················· | ···················· | ···················· | |
| AB | 280 | hCoV-19/England/QEUH-CAD90E/2020\|EPI_ISL_761239\|20 | ···················· | ···················· | ····················· | ···················· | ···················· | ···················· | |
| AB | 281 | hCoV-19/England/ALDP-CB3589/2020\|EPI_ISL_761240\|20 | ···················· | ···················· | ····················· | ···················· | ···················· | ···················· | |
| AB | 282 | hCoV-19/England/QEUH-CAD93B/2020\|EPI_ISL_761241\|20 | ···················· | ···················· | ····················· | ···················· | ···················· | ···················· | |
| AB | 283 | hCoV-19/England/ALDP-CB4DBD/2020\|EPI_ISL_761242\|20 | ···················· | ···················· | ····················· | ···················· | ···················· | ···················· | |
| AB | 284 | hCoV-19/England/ALDP-CB36C2/2020\|EPI_ISL_761243\|20 | ···················· | ···················· | ····················· | ···················· | ···················· | ···················· | |
| AB | 285 | hCoV-19/England/ALDP-CB4667/2020\|EPI_ISL_761244\|20 | ···················· | ···················· | ····················· | ···················· | ···················· | ···················· | |
| AB | 286 | hCoV-19/England/ALDP-CB46A3/2020\|EPI_ISL_761245\|20 | ···················· | ···················· | ····················· | ···················· | ···················· | ···················· | |
| AB | 287 | hCoV-19/England/ALDP-CB689C/2020\|EPI_ISL_761246\|20 | ···················· | ···················· | ····················· | ···················· | ···················· | ···················· | |
| AB | 288 | hCoV-19/England/ALDP-CB34D7/2020\|EPI_ISL_761247\|20 | ···················· | ···················· | ····················· | ···················· | ···················· | ···················· | |
| AB | 289 | hCoV-19/England/ALDP-CB3DEB/2020\|EPI_ISL_761248\|20 | ···················· | ···················· | ····················· | ···················· | ···················· | ···················· | |
| AB | 290 | hCoV-19/England/QEUH-CADEC3/2020\|EPI_ISL_761252\|20 | ···················· | ···················· | ····················· | ···················· | ···················· | ···················· | |
| AB | 291 | hCoV-19/England/ALDP-CB4694/2020\|EPI_ISL_761253\|20 | ···················· | ···················· | ····················· | ···················· | ···················· | ···················· | |
| AB | 292 | hCoV-19/England/ALDP-CB353E/2020\|EPI_ISL_761256\|20 | ···················· | ···················· | ····················· | ···················· | ···················· | ···················· | |
| AB | 293 | hCoV-19/England/QEUH-CADC9C/2020\|EPI_ISL_761259\|20 | ···················· | ···················· | ····················· | ···················· | ···················· | ···················· | |
| AB | 294 | hCoV-19/England/ALDP-CB5D43/2020\|EPI_ISL_761260\|20 | ···················· | ·A·················· | ····················· | ···················· | ···················· | ···A················ | |
| AB | 295 | hCoV-19/England/ALDP-CB4069/2020\|EPI_ISL_761261\|20 | ···················· | ···················· | ····················· | ···················· | ···················· | ···················· | |
| AB | 296 | hCoV-19/England/ALDP-CB4676/2020\|EPI_ISL_761262\|20 | ···················· | ···················· | ····················· | ···················· | ···················· | ···················· | |
| AB | 297 | hCoV-19/England/ALDP-CB48CB/2020\|EPI_ISL_761263\|20 | ···················· | ···················· | ····················· | ···················· | ···················· | ···················· | |
| AB | 298 | hCoV-19/England/ALDP-CB36A4/2020\|EPI_ISL_761264\|20 | ···················· | ···················· | ····················· | ···················· | ···················· | ···················· | |
| AB | 299 | hCoV-19/England/ALDP-CB4B59/2020\|EPI_ISL_761265\|20 | ···················· | ···················· | ····················· | ···················· | ···················· | ···················· | |
| AB | 300 | hCoV-19/England/ALDP-CB49D7/2020\|EPI_ISL_761270\|20 | ···················· | ···················· | ····················· | ···················· | ···················· | ···················· | |
| AB | 301 | hCoV-19/England/ALDP-CB3E51/2020\|EPI_ISL_761271\|20 | ···················· | ···················· | ····················· | ···················· | ···················· | ···················· | |
| AB | 302 | hCoV-19/England/ALDP-CB355C/2020\|EPI_ISL_761273\|20 | ···················· | ···················· | ····················· | ···················· | ···················· | ···················· | |
| AB | 303 | hCoV-19/England/ALDP-CB3ECA/2020\|EPI_ISL_761275\|20 | ···················· | ···················· | ····················· | ···················· | ···················· | ···················· | |
| AB | 304 | hCoV-19/England/QEUH-CAD494/2020\|EPI_ISL_761276\|20 | ···················· | ···················· | ····················· | ···················· | ······T············· | ···················· | |
| AB | 305 | hCoV-19/England/ALDP-CB4D44/2020\|EPI_ISL_761277\|20 | ···················· | ···················· | ····················· | ···················· | ···················· | ···················· | |
| AB | 306 | hCoV-19/England/ALDP-CB498C/2020\|EPI_ISL_761279\|20 | ···················· | ···················· | ····················· | ···················· | ···················· | ···················· | |
| AB | 307 | hCoV-19/England/ALDP-CB363B/2020\|EPI_ISL_761281\|20 | ···················· | ···················· | ····················· | ···················· | ···················· | ···················· | |
| AB | 308 | hCoV-19/England/ALDP-CB3FE5/2020\|EPI_ISL_761282\|20 | ···················· | ···················· | ····················· | ···················· | ···················· | ···················· | |
| AB | 309 | hCoV-19/England/QEUH-CADF75/2020\|EPI_ISL_761283\|20 | ···················· | ···················· | ····················· | ···················· | ···················· | ···················· | |

| | | | | | | | | |
|---|---|---|---|---|---|---|---|---|
| AB | 310 | hCoV-19/England/ALDP-CB3EBB/2020\|EPI_ISL_761284\|20 | ···················· | ···················· | ···················· | ···················· | ···················· | ···················· |
| AB | 311 | hCoV-19/England/ALDP-CB4DAE/2020\|EPI_ISL_761285\|20 | ···················· | ···················· | ···················· | ···················· | ···················· | ···················· |
| AB | 312 | hCoV-19/England/ALDP-CB34C8/2020\|EPI_ISL_761290\|20 | ···················· | ···················· | ···················· | ···················· | ···················· | ···················· |
| AB | 313 | hCoV-19/England/ALDP-CB5BD0/2020\|EPI_ISL_761291\|20 | ···················· | ···················· | ···················· | ···················· | ···················· | ···················· |
| AB | 314 | hCoV-19/England/ALDP-CB399C/2020\|EPI_ISL_761295\|20 | ···················· | ···················· | ···················· | ···················· | ···················· | ···················· |
| AB | 315 | hCoV-19/England/QEUH-CAD91D/2020\|EPI_ISL_761296\|20 | ···················· | ···················· | ···················· | ···················· | ···················· | ···················· |
| AB | 316 | hCoV-19/England/ALDP-CB39AB/2020\|EPI_ISL_761297\|20 | ···················· | ···················· | ···················· | ···················· | ···················· | ···················· |
| AB | 317 | hCoV-19/England/MILK-CA4B89/2020\|EPI_ISL_762502\|20 | ···················· | ···················· | ···················· | ···················· | ···················· | ···················· |
| AB | 318 | hCoV-19/England/MILK-CA9771/2020\|EPI_ISL_762503\|20 | ···················· | ···················· | ···················· | ···················· | ···················· | ···················· |
| AB | 319 | hCoV-19/England/MILK-CA9911/2020\|EPI_ISL_762504\|20 | ···················· | ···················· | ···················· | ···················· | ···················· | ···················· |
| AB | 320 | hCoV-19/England/MILK-CA9D06/2020\|EPI_ISL_762507\|20 | ···················· | ···················· | ···················· | ···················· | ···················· | ···················· |
| AB | 321 | hCoV-19/England/MILK-CA9D7F/2020\|EPI_ISL_762510\|20 | ···················· | ···················· | ···················· | ···················· | ···················· | ···················· |
| AB | 322 | hCoV-19/England/MILK-CA4D56/2020\|EPI_ISL_762511\|20 | ···················· | ···················· | ···················· | ···················· | ···················· | ···················· |
| AB | 323 | hCoV-19/England/MILK-CA9C72/2020\|EPI_ISL_762518\|20 | ···················· | ···················· | ···················· | ···················· | ···················· | ···················· |
| AB | 324 | hCoV-19/England/MILK-CA45B8/2020\|EPI_ISL_762521\|20 | ···················· | ···················· | ···················· | ···················· | ···················· | ···················· |
| AB | 325 | hCoV-19/England/MILK-CA4B10/2020\|EPI_ISL_762523\|20 | ···················· | ···················· | ···················· | ···················· | ···················· | ···················· |
| AB | 326 | hCoV-19/England/MILK-CA9BDF/2020\|EPI_ISL_762524\|20 | ···················· | ···················· | ···················· | ···················· | ···················· | ···················· |
| AB | 327 | hCoV-19/England/MILK-CA937D/2020\|EPI_ISL_762539\|20 | ···················· | ···················· | ···················· | ···················· | ···················· | ···················· |
| AB | 328 | hCoV-19/England/MILK-CA458B/2020\|EPI_ISL_762542\|20 | ···················· | ···················· | ···················· | ···················· | ···················· | ···················· |
| AB | 329 | hCoV-19/England/MILK-CA936E/2020\|EPI_ISL_762543\|20 | ···················· | ···················· | ···················· | ···················· | ···················· | ···················· |
| AB | 330 | hCoV-19/England/CAMC-C8391D/2020\|EPI_ISL_762549\|20 | ···················· | ···················· | ···················· | ···················· | ···················· | ···················· |
| AB | 331 | hCoV-19/England/ALDP-C88A06/2020\|EPI_ISL_762558\|20 | ···················· | ···················· | ···················· | ···················· | ·····T·············· | ···················· |
| AB | 332 | hCoV-19/England/ALDP-C88426/2020\|EPI_ISL_762562\|20 | ···················· | ···················· | ···················· | ···················· | ···················· | ···················· |
| AB | 333 | hCoV-19/England/CAMC-C804F1/2020\|EPI_ISL_762563\|20 | ···················· | ···················· | ···················· | ···················· | ···················· | ···················· |
| AB | 334 | hCoV-19/England/CAMC-C807CB/2020\|EPI_ISL_762572\|20 | ···················· | ···················· | ···················· | ···················· | ···················· | ···················· |
| AB | 335 | hCoV-19/England/CAMC-C836DA/2020\|EPI_ISL_762580\|20 | ···················· | ···················· | ···················· | ···················· | ·····T·············· | ···················· |
| AB | 336 | hCoV-19/England/QEUH-C7C6D4/2020\|EPI_ISL_762586\|20 | ···················· | ···················· | ···················· | ···················· | ···················· | ···················· |
| AB | 338 | hCoV-19/USA/CA-LACPHL-AF00073/2020\|EPI_ISL_766010\| | ···················· | ·A·················· | ···················· | ···················· | ···················· | ···················· |
| AB | 339 | hCoV-19/USA/CA-LACPHL-AF00074/2020\|EPI_ISL_766011\| | ···················· | ···················· | ···················· | ···················· | ···················· | ···················· |
| AB | 340 | hCoV-19/USA/CA-LACPHL-AF00075/2020\|EPI_ISL_766012\| | ···················· | ·A·················· | ···················· | ···················· | ···················· | ···················· |
| AB | 341 | hCoV-19/USA/CA-LACPHL-AF00076/2020\|EPI_ISL_766013\| | ···················· | ···················· | ···················· | ···················· | ···················· | ···················· |
| AB | 342 | hCoV-19/USA/CA-LACPHL-AF00077/2020\|EPI_ISL_766014\| | ···················· | ···················· | ···················· | ···················· | ···················· | ···················· |
| AB | 343 | hCoV-19/USA/CA-LACPHL-AE00056/2020\|EPI_ISL_766015\| | ···················· | ···················· | ···················· | ···················· | ···················· | ···················· |
| AB | 344 | hCoV-19/Vietnam/40139/2020\|EPI_ISL_766029\|2020-07 | ···················· | ···················· | ···················· | ···················· | ···················· | ···················· |
| AB | 345 | hCoV-19/Indonesia/YO-EIJK08/2020\|EPI_ISL_766032\|20 | ···················· | ···················· | ···················· | ···················· | ···················· | ···················· |
| AB | 346 | hCoV-19/Indonesia/NT-EIJK14/2020\|EPI_ISL_766033\|20 | ···················· | ···················· | ···················· | ···················· | ···················· | ···················· |
| AB | 347 | hCoV-19/Indonesia/BT-EIJK17/2020\|EPI_ISL_766034\|20 | ···················· | ···················· | ···················· | ···················· | ···················· | ···················· |
| AB | 348 | hCoV-19/Indonesia/JB-EIJK18/2020\|EPI_ISL_766035\|20 | ···················· | ···················· | ···················· | ···················· | ···················· | ···················· |
| AB | 349 | hCoV-19/Indonesia/BT-EIJK21/2020\|EPI_ISL_766036\|20 | ···················· | ···················· | ···················· | ···················· | ···················· | ···················· |
| AB | 350 | hCoV-19/Indonesia/JK-EIJK22/2020\|EPI_ISL_766037\|20 | ···················· | ···················· | ···················· | ···················· | ···················· | ···················· |
| AB | 351 | hCoV-19/Indonesia/JK-EIJK25/2020\|EPI_ISL_766038\|20 | ···················· | ···················· | ···················· | ···················· | ···················· | ···················· |
| AB | 352 | hCoV-19/Indonesia/NT-EIJK27/2020\|EPI_ISL_766039\|20 | ···················· | ···················· | ···················· | ···················· | ···················· | ···················· |
| AB | 353 | hCoV-19/Indonesia/JK-EIJK28/2020\|EPI_ISL_766040\|20 | ···················· | ···················· | ···················· | ···················· | ···················· | ···················· |
| AB | 354 | hCoV-19/Indonesia/JK-EIJK29/2020\|EPI_ISL_766041\|20 | ···················· | ···················· | ···················· | ···················· | ···················· | ···················· |
| AB | 355 | hCoV-19/Indonesia/BT-EIJK30/2020\|EPI_ISL_766042\|20 | ···················· | ···················· | ···················· | ···················· | ···················· | ···················· |
| AB | 356 | hCoV-19/Indonesia/JK-EIJK31/2020\|EPI_ISL_766043\|20 | ···················· | ···················· | ···················· | ···················· | ···················· | ···················· |
| AB | 357 | hCoV-19/Indonesia/JK-EIJK33/2020\|EPI_ISL_766044\|20 | ···················· | ···················· | ···················· | ···················· | ···················· | ···················· |
| AB | 358 | hCoV-19/Indonesia/JK-EIJK35/2020\|EPI_ISL_766045\|20 | ···················· | ···················· | ···················· | ···················· | ···················· | ···················· |
| AB | 359 | hCoV-19/Indonesia/JK-EIJK37/2020\|EPI_ISL_766046\|20 | ···················· | ···················· | ···················· | ···················· | ···················· | ···················· |
| AB | 360 | hCoV-19/Indonesia/JK-EIJK44/2020\|EPI_ISL_766047\|20 | ···················· | ···················· | ···················· | ···················· | ···················· | ···················· |
| AB | 361 | hCoV-19/Indonesia/NT-EIJK26/2020\|EPI_ISL_766048\|20 | ···················· | ···················· | ···················· | ···················· | ···················· | ···················· |
| AB | 362 | hCoV-19/USA/NY-Wadsworth-225541-01/2020\|EPI_ISL_76 | ···················· | ···················· | ···················· | ···················· | ···················· | ···················· |
| AB | 363 | hCoV-19/Australia/NSW1384/2020\|EPI_ISL_768606\|2020 | ···················· | ···················· | ···················· | ···················· | ···················· | ···················· |
| AB | 364 | hCoV-19/Australia/NSW1385/2020\|EPI_ISL_768607\|2020 | ···················· | ···················· | ···················· | ···················· | ···················· | ···················· |
| AB | 365 | hCoV-19/Thailand/Chiang Rai_MM/2020\|EPI_ISL_768609 | ···················· | ·A·················· | ···················· | ···················· | ···················· | ··A················· |
| AB | 366 | hCoV-19/Thailand/Chonburi_PN0160/2020\|EPI_ISL_7686 | ···················· | ···················· | ···················· | ···················· | ···················· | ···················· |
| AB | 367 | hCoV-19/Singapore/1503/2020\|EPI_ISL_768619\|2020-12 | ···················· | ···················· | ···················· | ···················· | ···················· | ···················· |
| AB | 368 | hCoV-19/Singapore/1504/2020\|EPI_ISL_768620\|2020-12 | ···················· | ···················· | ···················· | ···················· | ···················· | ···················· |
| AB | 369 | hCoV-19/Singapore/1505/2020\|EPI_ISL_768621\|2020-12 | ···················· | ···················· | ···················· | ···················· | ···················· | ··A················· |
| AB | 370 | hCoV-19/Singapore/1506/2020\|EPI_ISL_768622\|2020-12 | ···················· | ···················· | ···················· | ···················· | ···················· | ···················· |
| AB | 371 | hCoV-19/Singapore/1507/2020\|EPI_ISL_768623\|2020-12 | ···················· | ···················· | ···················· | ···················· | ···················· | ···················· |
| AB | 372 | hCoV-19/Singapore/1508/2020\|EPI_ISL_768624\|2020-12 | ···················· | ···················· | ···················· | ···················· | ···················· | ··A················· |
| AB | 373 | hCoV-19/Singapore/1509/2020\|EPI_ISL_768625\|2020-12 | ···················· | ···················· | ···················· | ···················· | ···················· | ···················· |
| AB | 374 | hCoV-19/Singapore/1510/2020\|EPI_ISL_768626\|2020-12 | ···················· | ···················· | ···················· | ···················· | ···················· | ···················· |
| AB | 375 | hCoV-19/Japan/IC-0461/2020\|EPI_ISL_768670\|2020-12 | ···················· | ···················· | ···················· | ···················· | ···················· | ···················· |
| AB | 376 | hCoV-19/Japan/IC-0462/2020\|EPI_ISL_768671\|2020-12 | ···················· | ···················· | ···················· | ···················· | ···T················ | ···················· |
| AB | 377 | hCoV-19/Japan/IC-0463/2020\|EPI_ISL_768672\|2020-12 | ···················· | ·A·················· | ···················· | ···················· | ···················· | ···················· |
| AB | 378 | hCoV-19/Japan/IC-0464/2020\|EPI_ISL_768673\|2020-12 | ···················· | ·A·················· | ···················· | ···················· | ···················· | ···················· |
| AB | 379 | hCoV-19/Japan/IC-0465/2020\|EPI_ISL_768674\|2020-12 | ···················· | ···················· | ···················· | ···················· | ···················· | ···················· |
| AB | 380 | hCoV-19/Japan/IC-0466/2020\|EPI_ISL_768675\|2020-12 | ···················· | ···················· | ···················· | ···················· | ···················· | ···················· |
| AB | 381 | hCoV-19/Japan/IC-0468/2020\|EPI_ISL_768677\|2020-12 | ···················· | ···················· | ···················· | ···················· | ···················· | ···················· |
| AB | 382 | hCoV-19/Japan/IC-0469/2020\|EPI_ISL_768678\|2020-12 | ···················· | ···················· | ···················· | ···················· | ···················· | ···················· |
| AB | 383 | hCoV-19/Japan/IC-0470/2020\|EPI_ISL_768679\|2020-12 | ···················· | ···················· | ···················· | ···················· | ···················· | ···················· |
| AB | 384 | hCoV-19/Japan/IC-0471/2020\|EPI_ISL_768680\|2020-12 | ···················· | ···················· | ···················· | ···················· | ···················· | ···················· |
| AB | 385 | hCoV-19/Japan/IC-0472/2020\|EPI_ISL_768681\|2020-12 | ···················· | ···················· | ···················· | ···················· | ···················· | ···················· |
| AB | 386 | hCoV-19/Japan/IC-0473/2020\|EPI_ISL_768682\|2020-12 | ···················· | ···················· | ···················· | ···················· | ···················· | ···················· |
| AB | 387 | hCoV-19/Japan/IC-0474/2020\|EPI_ISL_768683\|2020-12 | ···················· | ···················· | ···················· | ···················· | ···················· | ···················· |
| AB | 388 | hCoV-19/Japan/IC-0475/2020\|EPI_ISL_768684\|2020-12 | ···················· | ·A·················· | ···················· | ···················· | ···················· | ··A················· |
| AB | 389 | hCoV-19/Japan/IC-0476/2020\|EPI_ISL_768685\|2020-12 | ···················· | ···················· | ···················· | ···················· | ···················· | ···················· |
| AB | 390 | hCoV-19/Japan/IC-0477/2020\|EPI_ISL_768686\|2020-12 | ···················· | ···················· | ···················· | ···················· | ···················· | ···················· |

```
AB    391 hCoV-19/Japan/IC-0479/2020|EPI_ISL_768688|2020-12    ····················  ····················  ····················  ····················  ····················  ····················
AB    392 hCoV-19/Japan/IC-0480/2020|EPI_ISL_768689|2020-12    ····················  ····················  ····················  ····················  ····················  ····················
AB    393 hCoV-19/Japan/IC-0482/2020|EPI_ISL_768691|2020-12    ····················  ·A··················  ····················  ····················  ····················  ··A·················
AB    394 hCoV-19/Japan/IC-0483/2020|EPI_ISL_768692|2020-12    ····················  ·A··················  ····················  ····················  ····················  ··A·················
AB    395 hCoV-19/Japan/IC-0484/2020|EPI_ISL_768693|2020-12    ····················  ····················  ····················  ····················  ····················  ····················
AB    396 hCoV-19/Japan/IC-0487/2020|EPI_ISL_768696|2020-12    ········T···········  ····················  ····················  ····················  ····················  ····················
AB    397 hCoV-19/Japan/IC-0488/2020|EPI_ISL_768697|2020-12    ········T···········  ····················  ····················  ····················  ····················  ····················
AB    398 hCoV-19/Japan/IC-0489/2020|EPI_ISL_768698|2020-12    ····················  ·A··················  ····················  ····················  ····················  ····················
AB    399 hCoV-19/Japan/IC-0490/2020|EPI_ISL_768699|2020-12    ····················  ····················  ····················  ····················  ····················  ····················
AB    400 hCoV-19/Japan/IC-0491/2020|EPI_ISL_768700|2020-12    ····················  ····················  ····················  ····················  ····················  ····················
AB    401 hCoV-19/Japan/IC-0492/2020|EPI_ISL_768701|2020-12    ····················  ····················  ····················  ····················  ····················  ····················
AB    402 hCoV-19/Japan/IC-0493/2020|EPI_ISL_768702|2020-11    ····················  ····················  ····················  ····················  ····················  ····················
AB    403 hCoV-19/Japan/IC-0494/2020|EPI_ISL_768703|2020-12    ····················  ····················  ····················  ····················  ····················  ····················
AB    404 hCoV-19/Ireland/D-NVRL-AIIDV0073v1/2020|EPI_ISL_76    ····················  ····················  ····················  ····················  ····················  ····················
AB    405 hCoV-19/USA/NY-Wadsworth-292746-01/2020|EPI_ISL_76    ····················  ····················  ····················  ····················  ····················  ····················
AB    406 hCoV-19/USA/NY-Wadsworth-292748-01/2020|EPI_ISL_76    ····················  ····················  ····················  ····················  ····················  ····················
AB    407 hCoV-19/USA/NY-Wadsworth-292749-01/2020|EPI_ISL_76    ····················  ·A··················  ····················  ····················  ····················  ····················
AB    408 hCoV-19/USA/NY-Wadsworth-292750-01/2020|EPI_ISL_76    ····················  ·A··················  ····················  ····················  ····················  ····················
AB    409 hCoV-19/USA/NY-Wadsworth-292751-01/2020|EPI_ISL_76    ····················  ····················  ····················  ····················  ····················  ····················
AB    410 hCoV-19/USA/NY-Wadsworth-292760-01/2020|EPI_ISL_76    ····················  ····················  ····················  ····················  ····················  ····················
AB    411 hCoV-19/USA/NY-Wadsworth-292761-01/2020|EPI_ISL_76    ····················  ····················  ····················  ····················  ····················  ····················
AB    412 hCoV-19/USA/NY-Wadsworth-292762-01/2020|EPI_ISL_76    ····················  ·A··················  ····················  ····················  ····················  ····················
AB    413 hCoV-19/USA/NY-Wadsworth-292764-01/2020|EPI_ISL_76    ····················  ·A··················  ····················  ····················  ····················  ····················
AB    414 hCoV-19/USA/NY-Wadsworth-292765-01/2020|EPI_ISL_76    ····················  ····················  ····················  ····················  ····················  ····················
AB    415 hCoV-19/USA/NY-Wadsworth-292768-01/2020|EPI_ISL_76    ····················  ····················  ····················  ····················  ····················  ····················
AB    416 hCoV-19/USA/NY-Wadsworth-292769-01/2020|EPI_ISL_76    ····················  ····················  ····················  ····················  ····················  ····················
AB    417 hCoV-19/USA/NY-Wadsworth-292771-01/2020|EPI_ISL_76    ····················  ·A··················  ····················  ····················  ····················  ····················
AB    418 hCoV-19/USA/NY-Wadsworth-292772-01/2020|EPI_ISL_76    ····················  ·A··················  ····················  ····················  ····················  ····················
AB    419 hCoV-19/USA/NY-Wadsworth-292774-01/2020|EPI_ISL_76    ····················  ·A··················  ····················  ····················  ····················  ····················
AB    420 hCoV-19/USA/NY-Wadsworth-292775-01/2020|EPI_ISL_76    ····················  ·A··················  ····················  ····················  ····················  ····················
AB    421 hCoV-19/USA/NY-Wadsworth-292778-01/2020|EPI_ISL_76    ····················  ····················  ····················  ····················  ····················  ····················
AB    422 hCoV-19/USA/NY-Wadsworth-292779-01/2020|EPI_ISL_76    ····················  ·A··················  ····················  ····················  ····················  ····················
AB    423 hCoV-19/USA/NY-Wadsworth-292780-01/2020|EPI_ISL_76    ····················  ·A··················  ····················  ····················  ····················  ····················
AB    424 hCoV-19/USA/NY-Wadsworth-292783-01/2020|EPI_ISL_76    ····················  ····················  ····················  ····················  ····················  ····················
AB    425 hCoV-19/USA/NY-Wadsworth-292784-01/2020|EPI_ISL_76    ····················  ····················  ····················  ····················  ····················  ····················
AB    426 hCoV-19/USA/NY-Wadsworth-292785-01/2020|EPI_ISL_76    ·················T··  ····················  ····················  ····················  ····················  ····················    1
AB    427 hCoV-19/USA/NY-Wadsworth-292786-01/2020|EPI_ISL_76    ····················  ····················  ····················  ····················  ····················  ····················
AB    428 hCoV-19/USA/NY-Wadsworth-292787-01/2020|EPI_ISL_76    ····················  ····················  ····················  ····················  ····················  ····················
AB    429 hCoV-19/USA/NY-Wadsworth-292788-01/2020|EPI_ISL_76    ····················  ·A··················  ····················  ····················  ····················  ····················
AB    430 hCoV-19/USA/NY-Wadsworth-292790-01/2020|EPI_ISL_76    ····················  ····················  ····················  ····················  ····················  ····················
AB    431 hCoV-19/USA/NY-Wadsworth-292791-01/2020|EPI_ISL_76    ····················  ····················  ····················  ····················  ····················  ····················
AB    432 hCoV-19/Luxembourg/LNS0445708/2020|EPI_ISL_770905|    ····················  ····················  ····················  ····················  ····················  ····················
AB    433 hCoV-19/Luxembourg/LNS1999667/2020|EPI_ISL_770906|    ····················  ····················  ····················  ····················  ····················  ····················
AB    434 hCoV-19/Luxembourg/LNS3688196/2020|EPI_ISL_770907|    ····················  ····················  ····················  ····················  ····················  ····················
AB    435 hCoV-19/Luxembourg/LNS4812092/2020|EPI_ISL_770908|    ····················  ····················  ····················  ····················  ····················  ····················
AB    436 hCoV-19/Luxembourg/LNS7445177/2020|EPI_ISL_770909|    ····················  ····················  ····················  ····················  ····················  ····················
AB    437 hCoV-19/Luxembourg/LNS5107725/2020|EPI_ISL_770920|    ····················  ····················  ····················  ····················  ····················  ····················
AB    438 hCoV-19/Luxembourg/LNS6370076/2020|EPI_ISL_770921|    ····················  ····················  ····················  ····················  ····················  ····················
AB    439 hCoV-19/Luxembourg/LNS6087810/2020|EPI_ISL_770922|    ····················  ····················  ····················  ····················  ····················  ····················
AB    440 hCoV-19/Luxembourg/LNS8835096/2020|EPI_ISL_770923|    ····················  ····················  ····················  ····················  ····················  ····················
AB    441 hCoV-19/Luxembourg/LNS9974678/2020|EPI_ISL_770924|    ····················  ····················  ····················  ····················  ····················  ····················
AB    442 hCoV-19/Luxembourg/LNS8348160/2020|EPI_ISL_770925|    ····················  ····················  ····················  ····················  ····················  ····················
AB    443 hCoV-19/Luxembourg/LNS5966742/2020|EPI_ISL_770926|    ····················  ····················  ····················  ····················  ····················  ····················
AB    444 hCoV-19/Luxembourg/LNS2162625/2020|EPI_ISL_770927|    ····················  ····················  ····················  ····················  ····················  ····················
AB    445 hCoV-19/Luxembourg/LNS2883051/2020|EPI_ISL_770928|    ····················  ····················  ····················  ····················  ····················  ····················
AB    446 hCoV-19/Luxembourg/LNS4090457/2020|EPI_ISL_770929|    ····················  ····················  ····················  ····················  ····················  ····················
AB    447 hCoV-19/Luxembourg/LNS3537918/2020|EPI_ISL_770930|    ····················  ····················  ····················  ····················  ····················  ····················
AB    448 hCoV-19/Luxembourg/LNS4596559/2020|EPI_ISL_770931|    ····················  ····················  ····················  ····················  ····················  ····················
AB    449 hCoV-19/Luxembourg/LNS1448912/2020|EPI_ISL_770932|    ····················  ···A················  ····················  ····················  ····················  ····················
AB    450 hCoV-19/Luxembourg/LNS3477613/2020|EPI_ISL_770933|    ····················  ····················  ····················  ····················  ·····T··············  ····················
AB    451 hCoV-19/Luxembourg/LNS5453199/2020|EPI_ISL_770934|    ····················  ····················  ····················  ····················  ····················  ····················
AB    452 hCoV-19/Luxembourg/LNS1033433/2020|EPI_ISL_770935|    ····················  ····················  ····················  ····················  ····················  ····················
AB    453 hCoV-19/Luxembourg/LNS2609501/2020|EPI_ISL_770936|    ····················  ····················  ····················  ····················  ····················  ····················
AB    454 hCoV-19/Luxembourg/LNS8720271/2020|EPI_ISL_770937|    ····················  ····················  ····················  ····················  ····················  ····················
AB    455 hCoV-19/Luxembourg/LNS7935054/2020|EPI_ISL_770938|    ····················  ····················  ····················  ····················  ····················  ····················
AB    456 hCoV-19/Luxembourg/LNS9113369/2020|EPI_ISL_770939|    ····················  ····················  ····················  ····················  ····················  ····················
AB    457 hCoV-19/Luxembourg/LNS0276550/2020|EPI_ISL_770981|    ····················  ····················  ····················  ····················  ····················  ····················
AB    458 hCoV-19/Luxembourg/LNS4800679/2020|EPI_ISL_770982|    ····················  ····················  ····················  ····················  ····················  ····················
AB    459 hCoV-19/Luxembourg/LNS2320968/2020|EPI_ISL_770983|    ····················  ····················  ····················  ····················  ····················  ····················
AB    460 hCoV-19/Luxembourg/LNS7071419/2020|EPI_ISL_770984|    ····················  ····················  ····················  ····················  ····················  ····················
AB    461 hCoV-19/Luxembourg/LNS1035636/2020|EPI_ISL_770985|    ····················  ····················  ····················  ····················  ····················  ····················
AB    462 hCoV-19/Luxembourg/LNS5529043/2020|EPI_ISL_770986|    ····················  ····················  ····················  ····················  ····················  ····················
AB    463 hCoV-19/Luxembourg/LNS8729762/2020|EPI_ISL_770987|    ····················  ····················  ····················  ····················  ····················  ····················
AB    464 hCoV-19/Luxembourg/LNS2786709/2020|EPI_ISL_770988|    ····················  ····················  ····················  ····················  ····················  ····················
AB    465 hCoV-19/Luxembourg/LNS6084294/2020|EPI_ISL_770989|    ····················  ····················  ····················  ····················  ····················  ····················
AB    466 hCoV-19/Luxembourg/LNS1247055/2020|EPI_ISL_770990|    ····················  ····················  ····················  ····················  ····················  ····················
AB    467 hCoV-19/Luxembourg/LNS4918054/2020|EPI_ISL_770991|    ····················  ····················  ····················  ····················  ····················  ····················
AB    468 hCoV-19/Luxembourg/LNS9293666/2020|EPI_ISL_770992|    ····················  ····················  ····················  ····················  ····················  ····················
AB    469 hCoV-19/Luxembourg/LNS4511488/2020|EPI_ISL_770993|    ····················  ····················  ····················  ····················  ····················  ····················
AB    470 hCoV-19/Luxembourg/LNS7075526/2020|EPI_ISL_770994|    ····················  ····················  ····················  ····················  ····················  ····················
```

| | | | | | | | | |
|---|---|---|---|---|---|---|---|---|
| AB | 471 | hCoV-19/Luxembourg/LNS2575316/2020\|EPI_ISL_770995\| | ···················· | ···················· | ····················· | ···················· | ···················· | ···················· |
| AB | 472 | hCoV-19/Luxembourg/LNS8199676/2020\|EPI_ISL_770996\| | ···················· | ···················· | ····················· | ···················· | ···················· | ···················· |
| AB | 473 | hCoV-19/Luxembourg/LNS0298658/2020\|EPI_ISL_770997\| | ···················· | ···················· | ····················· | ···················· | ···················· | ···················· |
| AB | 474 | hCoV-19/Luxembourg/LNS6725409/2020\|EPI_ISL_770998\| | ···················· | ···················· | ····················· | ···················· | ···················· | ···················· |
| AB | 475 | hCoV-19/Luxembourg/LNS4041040/2020\|EPI_ISL_770999\| | ···················· | ···················· | ····················· | ···················· | ···················· | ···················· |
| AB | 476 | hCoV-19/England/QEUH-CC245D/2020\|EPI_ISL_778007\|20 | ···················· | ···················· | ····················· | ···················· | ···················· | ···················· |
| AB | 477 | hCoV-19/England/QEUH-CC3076/2020\|EPI_ISL_778014\|20 | ···················· | ···················· | ····················· | ···················· | ···················· | ···················· |
| AB | 478 | hCoV-19/England/CAMC-CB734F/2020\|EPI_ISL_778017\|20 | ···················· | ···················· | ····················· | ···················· | ···················· | ···················· |
| AB | 479 | hCoV-19/England/ALDP-CBF435/2020\|EPI_ISL_778018\|20 | ···················· | ···················· | ····················· | ···················· | ···················· | ···················· |
| AB | 480 | hCoV-19/England/CAMC-CB8FC2/2020\|EPI_ISL_778023\|20 | ···················· | ···················· | ····················· | ···················· | ···················· | ···················· |
| AB | 481 | hCoV-19/England/CAMC-CB6F00/2020\|EPI_ISL_778025\|20 | ···················· | ···················· | ····················· | ···················· | ···················· | ···················· |
| AB | 482 | hCoV-19/England/QEUH-CC2411/2020\|EPI_ISL_778026\|20 | ···················· | ···················· | ····················· | ···················· | ···················· | ···················· |
| AB | 483 | hCoV-19/England/CAMC-CB728E/2020\|EPI_ISL_778030\|20 | ···················· | ···················· | ····················· | ···················· | ···················· | ···················· |
| AB | 484 | hCoV-19/England/ALDP-CBF20E/2020\|EPI_ISL_778033\|20 | ···················· | ···················· | ····················· | ···················· | ···················· | ···················· |
| AB | 485 | hCoV-19/England/QEUH-CC3049/2020\|EPI_ISL_778034\|20 | ···················· | ···················· | ····················· | ···················· | ······T············· | ···················· |
| AB | 486 | hCoV-19/England/ALDP-CC097E/2020\|EPI_ISL_778035\|20 | ···················· | ···················· | ····················· | ···················· | ···················· | ···················· |
| AB | 487 | hCoV-19/England/CAMC-CB73D6/2020\|EPI_ISL_778037\|20 | ···················· | ···················· | ····················· | ···················· | ···················· | ···················· |
| AB | 488 | hCoV-19/England/QEUH-CC2D43/2020\|EPI_ISL_778042\|20 | ···················· | ···················· | ····················· | ···················· | ···················· | ···················· |
| AB | 489 | hCoV-19/England/ALDP-CBF9CD/2020\|EPI_ISL_778043\|20 | ···················· | ···················· | ····················· | ···················· | ···················· | ···················· |
| AB | 490 | hCoV-19/England/ALDP-CBF444/2020\|EPI_ISL_778044\|20 | ···················· | ···················· | ····················· | ···················· | ···················· | ···················· |
| AB | 491 | hCoV-19/England/CAMC-CB750D/2020\|EPI_ISL_778045\|20 | ···················· | ···················· | ····················· | ···················· | ···················· | ···················· |
| AB | 492 | hCoV-19/England/ALDP-CC0756/2020\|EPI_ISL_778047\|20 | ···················· | ···················· | ····················· | ···················· | ···················· | ···················· |
| AB | 493 | hCoV-19/England/ALDP-CC07B0/2020\|EPI_ISL_778048\|20 | ···················· | ···················· | ····················· | ···················· | ···················· | ···················· |
| AB | 494 | hCoV-19/England/QEUH-CC229F/2020\|EPI_ISL_778053\|20 | ···················· | ···················· | ····················· | ···················· | ···················· | ···················· |
| AB | 495 | hCoV-19/England/QEUH-CC301C/2020\|EPI_ISL_778058\|20 | ···················· | ···················· | ····················· | ···················· | ···················· | ···················· |
| AB | 496 | hCoV-19/England/ALDP-CC0747/2020\|EPI_ISL_778059\|20 | ···················· | ···················· | ····················· | ···················· | ···················· | ···················· |
| AB | 497 | hCoV-19/England/QEUH-CC22BD/2020\|EPI_ISL_778060\|20 | ···················· | ···················· | ····················· | ···················· | ···················· | ···················· |
| AB | 498 | hCoV-19/England/QEUH-CC2D70/2020\|EPI_ISL_778062\|20 | ···················· | ···················· | ····················· | ···················· | ···················· | ···················· |
| AB | 499 | hCoV-19/England/ALDP-CBF4DB/2020\|EPI_ISL_778065\|20 | ···················· | ···················· | ····················· | ···················· | ···················· | ···················· |
| AB | 500 | hCoV-19/England/CAMC-CB8F1D/2020\|EPI_ISL_778066\|20 | ···················· | ···················· | ····················· | ···················· | ···················· | ···················· |
| AB | 501 | hCoV-19/England/QEUH-CC3665/2020\|EPI_ISL_778068\|20 | ···················· | ···················· | ····················· | ···················· | ···················· | ···················· |
| AB | 502 | hCoV-19/England/QEUH-CC244E/2020\|EPI_ISL_778073\|20 | ···················· | ···················· | ····················· | ···················· | ···················· | ···················· |
| AB | 503 | hCoV-19/England/ALDP-CBF9FA/2020\|EPI_ISL_778074\|20 | ···················· | ···················· | ····················· | ···················· | ···················· | ···················· |
| AB | 504 | hCoV-19/England/QEUH-CC2DF8/2020\|EPI_ISL_778076\|20 | ···················· | ·A·················· | ····················· | ···················· | ···················· | ···A················ |
| AB | 505 | hCoV-19/England/QEUH-CC260C/2020\|EPI_ISL_778077\|20 | ···················· | ···················· | ····················· | ···················· | ···················· | ···················· |
| AB | 506 | hCoV-19/England/ALDP-CC0765/2020\|EPI_ISL_778083\|20 | ···················· | ···················· | ····················· | ···················· | ···················· | ···················· |
| AB | 507 | hCoV-19/England/CAMC-CB74C4/2020\|EPI_ISL_778085\|20 | ···················· | ···················· | ····················· | ···················· | ···················· | ···················· |
| AB | 508 | hCoV-19/England/ALDP-CBF471/2020\|EPI_ISL_778089\|20 | ···················· | ···················· | ····················· | ···················· | ·T·················· | ···················· |
| AB | 509 | hCoV-19/England/QEUH-CC27BE/2020\|EPI_ISL_778090\|20 | ···················· | ···················· | ····················· | ···················· | ···················· | ···A················ |
| AB | 510 | hCoV-19/England/QEUH-CC3674/2020\|EPI_ISL_778092\|20 | ···················· | ···················· | ····················· | ···················· | ···················· | ···················· |
| AB | 511 | hCoV-19/England/QEUH-CC25D2/2020\|EPI_ISL_778098\|20 | ···················· | ···················· | ····················· | ···················· | ···················· | ···················· |
| AB | 512 | hCoV-19/England/CAMC-CB7233/2020\|EPI_ISL_778099\|20 | ···················· | ···················· | ····················· | ···················· | ···················· | ···················· |
| AB | 513 | hCoV-19/South Korea/KDCA0235/2020\|EPI_ISL_760000\|2 | ···················· | ···················· | ····················· | ···················· | ···················· | ···················· |
| AB | 514 | hCoV-19/South Korea/KDCA0236/2020\|EPI_ISL_760001\|2 | ···················· | ···················· | ····················· | ···················· | ···················· | ···················· |
| AB | 515 | hCoV-19/South Korea/KDCA0237/2020\|EPI_ISL_760002\|2 | ···················· | ···················· | ····················· | ···················· | ···················· | ···················· |
| AB | 516 | hCoV-19/South Korea/KDCA0238/2020\|EPI_ISL_760003\|2 | ···················· | ···················· | ····················· | ···················· | ···················· | ···················· |
| AB | 517 | hCoV-19/South Korea/KDCA0239/2020\|EPI_ISL_760004\|2 | ···················· | ···················· | ····················· | ···················· | ···················· | ···················· |
| AB | 518 | hCoV-19/South Korea/KDCA0240/2020\|EPI_ISL_760005\|2 | ···················· | ···················· | ····················· | ···················· | ···················· | ···················· |
| AB | 519 | hCoV-19/South Korea/KDCA0241/2020\|EPI_ISL_760006\|2 | ···················· | ···················· | ····················· | ···················· | ···················· | ···················· |
| AB | 520 | hCoV-19/South Korea/KDCA0242/2020\|EPI_ISL_760007\|2 | ···················· | ···················· | ····················· | ···················· | ···················· | ···················· |
| AB | 521 | hCoV-19/South Korea/KDCA0244/2020\|EPI_ISL_760009\|2 | ···················· | ···················· | ····················· | ···················· | ···················· | ···················· |
| AB | 522 | hCoV-19/South Korea/KDCA0245/2020\|EPI_ISL_760010\|2 | ···················· | ···················· | ····················· | ···················· | ···················· | ···················· |
| AB | 523 | hCoV-19/South Korea/KDCA0246/2020\|EPI_ISL_760011\|2 | ···················· | ···················· | ····················· | ···················· | ···················· | ···················· |
| AB | 524 | hCoV-19/South Korea/KDCA0247/2020\|EPI_ISL_760012\|2 | ···················· | ···················· | ····················· | ···················· | ···················· | ···················· |
| AB | 525 | hCoV-19/South Korea/KDCA0248/2020\|EPI_ISL_760013\|2 | ···················· | ···················· | ····················· | ···················· | ···················· | ···················· |
| AB | 526 | hCoV-19/England/QEUH-CADBCC/2020\|EPI_ISL_761313\|20 | ···················· | ···················· | ····················· | ···················· | ···················· | ···················· |
| AB | 527 | hCoV-19/England/ALDP-CB3E33/2020\|EPI_ISL_761315\|20 | ···················· | ···················· | ····················· | ···················· | ···················· | ···················· |
| AB | 528 | hCoV-19/England/ALDP-CB34E6/2020\|EPI_ISL_761316\|20 | ···················· | ·········T·········· | ····················· | ···················· | ···················· | ···················· |
| AB | 529 | hCoV-19/England/ALDP-CB356B/2020\|EPI_ISL_761317\|20 | ···················· | ·········T·········· | ····················· | ···················· | ···················· | ···················· |
| AB | 530 | hCoV-19/England/ALDP-CB68F6/2020\|EPI_ISL_761318\|20 | ···················· | ···················· | ····················· | ···················· | ···················· | ···················· |
| AB | 531 | hCoV-19/England/ALDP-CB38AE/2020\|EPI_ISL_761319\|20 | ···················· | ···················· | ····················· | ···················· | ···················· | ···················· |
| AB | 532 | hCoV-19/England/ALDP-CB35E3/2020\|EPI_ISL_761320\|20 | ···················· | ···················· | ····················· | ···················· | ···················· | ···················· |
| AB | 533 | hCoV-19/England/ALDP-CB44B8/2020\|EPI_ISL_761321\|20 | ···················· | ···················· | ····················· | ···················· | ···················· | ···················· |
| AB | 534 | hCoV-19/England/ALDP-CB5D25/2020\|EPI_ISL_761322\|20 | ···················· | ·A·················· | ····················· | ···················· | ···················· | ···A················ |
| AB | 535 | hCoV-19/England/ALDP-CB679F/2020\|EPI_ISL_761323\|20 | ···················· | ···················· | ····················· | ···················· | ···················· | ···················· |
| AB | 536 | hCoV-19/England/ALDP-CB49B9/2020\|EPI_ISL_761328\|20 | ···················· | ···················· | ····················· | ···················· | ···················· | ···················· |
| AB | 537 | hCoV-19/England/ALDP-CB3E7F/2020\|EPI_ISL_761329\|20 | ···················· | ·A·················· | ····················· | ···················· | ···················· | ···················· |
| AB | 538 | hCoV-19/England/QEUH-CADC41/2020\|EPI_ISL_761331\|20 | ···················· | ···················· | ····················· | ···················· | ···················· | ···················· |
| AB | 539 | hCoV-19/England/ALDP-CB496E/2020\|EPI_ISL_761334\|20 | ···················· | ···················· | ····················· | ···················· | ···················· | ···················· |
| AB | 540 | hCoV-19/England/ALDP-CB4B4A/2020\|EPI_ISL_761335\|20 | ···················· | ···················· | ····················· | ···················· | ···················· | ···················· |
| AB | 541 | hCoV-19/England/ALDP-CB463A/2020\|EPI_ISL_761336\|20 | ···················· | ···················· | ····················· | ···················· | ···················· | ···················· |
| AB | 542 | hCoV-19/England/QEUH-CAD83E/2020\|EPI_ISL_761338\|20 | ···················· | ···················· | ····················· | ···················· | ···················· | ···················· |
| AB | 543 | hCoV-19/England/QEUH-CAD7C8/2020\|EPI_ISL_761339\|20 | ···················· | ···················· | ····················· | ···················· | ····T··············· | ···················· |
| AB | 544 | hCoV-19/England/ALDP-CB3DBE/2020\|EPI_ISL_761340\|20 | ···················· | ···················· | ····················· | ···················· | ···················· | ···················· |
| AB | 545 | hCoV-19/England/ALDP-CB67CC/2020\|EPI_ISL_761342\|20 | ···················· | ···················· | ····················· | ···················· | ···················· | ···················· |
| AB | 546 | hCoV-19/England/ALDP-CB360E/2020\|EPI_ISL_761344\|20 | ···················· | ···················· | ····················· | ···················· | ···················· | ···················· |
| AB | 547 | hCoV-19/Wales/ALDP-C8A309/2020\|EPI_ISL_762643\|2020 | ···················· | ···················· | ····················· | ···················· | ···················· | ···················· |
| AB | 548 | hCoV-19/England/ALDP-C88347/2020\|EPI_ISL_762647\|20 | ···················· | ···················· | ····················· | ···················· | ···················· | ···················· |
| AB | 549 | hCoV-19/England/QEUH-C7FE51/2020\|EPI_ISL_762648\|20 | ···················· | ···················· | ····················· | ···················· | ···················· | ···················· |
| AB | 550 | hCoV-19/England/QEUH-C7C689/2020\|EPI_ISL_762652\|20 | ···················· | ···················· | ····················· | ···················· | ···················· | ···················· |

| | | | | | | | | |
|---|---|---|---|---|---|---|---|---|
| AB | 551 | hCoV-19/England/ALDP-C8870F/2020\|EPI_ISL_762655\|20 | ···················· | ···················· | ···················· | ···················· | ···················· | ···················· |
| AB | 552 | hCoV-19/England/ALDP-C8A0A8/2020\|EPI_ISL_762659\|20 | ···················· | ···················· | ···················· | ···················· | ···················· | ···················· |
| AB | 553 | hCoV-19/England/ALDP-C889EB/2020\|EPI_ISL_762666\|20 | ···················· | ·A·················· | ···················· | ···················· | ···················· | ··A················· |
| AB | 554 | hCoV-19/England/CAMC-C81E3E/2020\|EPI_ISL_762667\|20 | ···················· | ···················· | ···················· | ···················· | ···················· | ···················· |
| AB | 556 | hCoV-19/USA/FL-BPHL-2265/2020\|EPI_ISL_767416\|2020- | ···················· | ···················· | ···················· | ···················· | ···················· | ···················· |
| AB | 557 | hCoV-19/USA/NY-Wadsworth-290724-01/2020\|EPI_ISL_76 | ···················· | ···················· | ···················· | ···················· | ···················· | ···················· |
| AB | 558 | hCoV-19/USA/NY-Wadsworth-287512-01/2020\|EPI_ISL_76 | ···················· | ···················· | ···················· | ···················· | ···················· | ···················· |
| AB | 559 | hCoV-19/USA/NY-Wadsworth-288932-01/2020\|EPI_ISL_76 | ···················· | ···················· | ···················· | ···················· | ···················· | ···················· |
| AB | 560 | hCoV-19/USA/NY-Wadsworth-290442-01/2020\|EPI_ISL_76 | ···················· | ···················· | ···················· | ···················· | ···················· | ···················· |
| AB | 561 | hCoV-19/USA/NY-Wadsworth-224452-01/2020\|EPI_ISL_76 | ···················· | ···················· | ···················· | ···················· | ···················· | ···················· |
| AB | 562 | hCoV-19/USA/NY-Wadsworth-224529-01/2020\|EPI_ISL_76 | ···················· | ···················· | ···················· | ···················· | ···················· | ···················· |
| AB | 563 | hCoV-19/USA/NY-Wadsworth-225796-01/2020\|EPI_ISL_76 | ···················· | ···················· | ···················· | ···················· | ···················· | ···················· |
| AB | 564 | hCoV-19/USA/NY-Wadsworth-225801-01/2020\|EPI_ISL_76 | ···················· | ···················· | ···················· | ···················· | ···················· | ···················· |
| AB | 565 | hCoV-19/USA/NY-Wadsworth-225865-01/2020\|EPI_ISL_76 | ···················· | ···················· | ···················· | ···················· | ···················· | ···················· |
| AB | 566 | hCoV-19/USA/NY-Wadsworth-226213-01/2020\|EPI_ISL_76 | ···················· | ···················· | ···················· | ···················· | ···················· | ···················· |
| AB | 567 | hCoV-19/USA/NY-Wadsworth-226222-01/2020\|EPI_ISL_76 | ···················· | ···················· | ···················· | ···················· | ···················· | ···················· |
| AB | 568 | hCoV-19/USA/NY-Wadsworth-226223-01/2020\|EPI_ISL_76 | ···················· | ···················· | ···················· | ···················· | ···················· | ···················· |
| AB | 569 | hCoV-19/USA/NY-Wadsworth-226226-01/2020\|EPI_ISL_76 | ···················· | ···················· | ···················· | ···················· | ···················· | ···················· |
| AB | 570 | hCoV-19/USA/NY-Wadsworth-226232-01/2020\|EPI_ISL_76 | ···················· | ···················· | ···················· | ···················· | ···················· | ···················· |
| AB | 571 | hCoV-19/USA/NY-Wadsworth-226378-01/2020\|EPI_ISL_76 | ···················· | ···················· | ···················· | ···················· | ···················· | ···················· |
| AB | 572 | hCoV-19/USA/NY-Wadsworth-226388-01/2020\|EPI_ISL_76 | ···················· | ···················· | ···················· | ···················· | ···················· | ···················· |
| AB | 573 | hCoV-19/USA/NY-Wadsworth-226389-01/2020\|EPI_ISL_76 | ···················· | ···················· | ···················· | ···················· | ···················· | ···················· |
| AB | 574 | hCoV-19/USA/NY-Wadsworth-228285-01/2020\|EPI_ISL_76 | ···················· | ···················· | ···················· | ···················· | ···················· | ···················· |
| AB | 575 | hCoV-19/USA/NY-Wadsworth-228302-01/2020\|EPI_ISL_76 | ···················· | ···················· | ···················· | ···················· | ···················· | ···················· |
| AB | 576 | hCoV-19/USA/NY-Wadsworth-228324-01/2020\|EPI_ISL_76 | ···················· | ···················· | ···················· | ···················· | ···················· | ···················· |
| AB | 577 | hCoV-19/USA/NY-Wadsworth-228345-01/2020\|EPI_ISL_76 | ···················· | ···················· | ···················· | ···················· | ···················· | ···················· |
| AB | 578 | hCoV-19/USA/NY-Wadsworth-228385-01/2020\|EPI_ISL_76 | ···················· | ···················· | ···················· | ···················· | ···················· | ···················· |
| AB | 579 | hCoV-19/USA/NY-Wadsworth-228418-01/2020\|EPI_ISL_76 | ···················· | ···················· | ···················· | ···················· | ···················· | ···················· |
| AB | 580 | hCoV-19/USA/NY-Wadsworth-228448-01/2020\|EPI_ISL_76 | ···················· | ···················· | ···················· | ···················· | ···················· | ···················· |
| AB | 581 | hCoV-19/USA/NY-Wadsworth-228529-01/2020\|EPI_ISL_76 | ···················· | ···················· | ···················· | ···················· | ···················· | ···················· |
| AB | 582 | hCoV-19/USA/NY-Wadsworth-229106-01/2020\|EPI_ISL_76 | ···················· | ···················· | ···················· | ···················· | ···················· | ···················· |
| AB | 583 | hCoV-19/USA/NY-Wadsworth-229107-01/2020\|EPI_ISL_76 | ···················· | ···················· | ···················· | ···················· | ···················· | ···················· |
| AB | 584 | hCoV-19/USA/NY-Wadsworth-229230-01/2020\|EPI_ISL_76 | ···················· | ···················· | ···················· | ···················· | ···················· | ···················· |
| AB | 585 | hCoV-19/USA/NY-Wadsworth-286664-01/2020\|EPI_ISL_76 | ···················· | ·A·················· | ···················· | ···················· | ···················· | ···················· |
| AB | 586 | hCoV-19/USA/NY-Wadsworth-286666-01/2020\|EPI_ISL_76 | ···················· | ···················· | ···················· | ···················· | ···················· | ···················· |
| AB | 587 | hCoV-19/USA/NY-Wadsworth-286673-01/2020\|EPI_ISL_76 | ···················· | ···················· | ···················· | ···················· | ···················· | ···················· |
| AB | 588 | hCoV-19/USA/NY-Wadsworth-286676-01/2020\|EPI_ISL_76 | ···················· | ···················· | ···················· | ···················· | ···················· | ···················· |
| AB | 589 | hCoV-19/USA/NY-Wadsworth-287341-01/2020\|EPI_ISL_76 | ···················· | ···················· | ···················· | ···················· | ···················· | ···················· |
| AB | 590 | hCoV-19/USA/NY-Wadsworth-287397-01/2020\|EPI_ISL_76 | ···················· | ···················· | ···················· | ···················· | ···················· | ···················· |
| AB | 591 | hCoV-19/USA/NY-Wadsworth-287401-01/2020\|EPI_ISL_76 | ···················· | ···················· | ···················· | ···················· | ···················· | ···················· |
| AB | 592 | hCoV-19/USA/NY-Wadsworth-287411-01/2020\|EPI_ISL_76 | ···················· | ·A·················· | ···················· | ···················· | ···················· | ···················· |
| AB | 593 | hCoV-19/USA/NY-Wadsworth-287412-01/2020\|EPI_ISL_76 | ···················· | ·A·················· | ···················· | ···················· | ···················· | ···················· |
| AB | 594 | hCoV-19/USA/NY-Wadsworth-287413-01/2020\|EPI_ISL_76 | ···················· | ·A·················· | ···················· | ···················· | ···················· | ···················· |
| AB | 595 | hCoV-19/USA/NY-Wadsworth-287417-01/2020\|EPI_ISL_76 | ···················· | ·A·················· | ···················· | ···················· | ···················· | ···················· |
| AB | 596 | hCoV-19/USA/NY-Wadsworth-287419-01/2020\|EPI_ISL_76 | ···················· | ···················· | ···················· | ···················· | ···················· | ···················· |
| AB | 597 | hCoV-19/USA/NY-Wadsworth-287495-01/2020\|EPI_ISL_76 | ···················· | ···················· | ···················· | ···················· | ···················· | ···················· |
| AB | 598 | hCoV-19/USA/NY-Wadsworth-287496-01/2020\|EPI_ISL_76 | ···················· | ···················· | ···················· | ···················· | ···················· | ···················· |
| AB | 599 | hCoV-19/USA/NY-Wadsworth-287498-01/2020\|EPI_ISL_76 | ···················· | ···················· | ···················· | ···················· | ···················· | ···················· |
| AB | 600 | hCoV-19/USA/NY-Wadsworth-287508-01/2020\|EPI_ISL_76 | ···················· | ·A·················· | ···················· | ···················· | ···················· | ···················· |
| AB | 601 | hCoV-19/USA/NY-Wadsworth-287513-01/2020\|EPI_ISL_76 | ···················· | ···················· | ···················· | ···················· | ···················· | ···················· |
| AB | 602 | hCoV-19/USA/NY-Wadsworth-287519-01/2020\|EPI_ISL_76 | ···················· | ···················· | ···················· | ···················· | ···················· | ···················· |
| AB | 603 | hCoV-19/USA/NY-Wadsworth-287521-01/2020\|EPI_ISL_76 | ···················· | ···················· | ···················· | ···················· | ···················· | ···················· |
| AB | 604 | hCoV-19/USA/NY-Wadsworth-287637-01/2020\|EPI_ISL_76 | ···················· | ···················· | ···················· | ···················· | ···················· | ···················· |
| AB | 605 | hCoV-19/USA/NY-Wadsworth-287696-01/2020\|EPI_ISL_76 | ···················· | ···················· | ···················· | ···················· | ···················· | ···················· |
| AB | 606 | hCoV-19/USA/NY-Wadsworth-287929-01/2020\|EPI_ISL_76 | ···················· | ·A·················· | ···················· | ···················· | ···················· | ···················· |
| AB | 607 | hCoV-19/USA/NY-Wadsworth-288163-01/2020\|EPI_ISL_76 | ···················· | ···················· | ···················· | ···················· | ···················· | ···················· |
| AB | 608 | hCoV-19/USA/NY-Wadsworth-288164-01/2020\|EPI_ISL_76 | ···················· | ·A·················· | ···················· | ···················· | ···················· | ···················· |
| AB | 609 | hCoV-19/USA/NY-Wadsworth-288186-01/2020\|EPI_ISL_76 | ···················· | ···················· | ···················· | ···················· | ···················· | ···················· |
| AB | 610 | hCoV-19/USA/NY-Wadsworth-288188-01/2020\|EPI_ISL_76 | ···················· | ···················· | ···················· | ···················· | ···················· | ···················· |
| AB | 611 | hCoV-19/USA/NY-Wadsworth-288189-01/2020\|EPI_ISL_76 | ···················· | ···················· | ···················· | ···················· | ···················· | ···················· |
| AB | 612 | hCoV-19/USA/NY-Wadsworth-288190-01/2020\|EPI_ISL_76 | ···················· | ···················· | ···················· | ···················· | ···················· | ···················· |
| AB | 613 | hCoV-19/USA/NY-Wadsworth-288191-01/2020\|EPI_ISL_76 | ···················· | ···················· | ···················· | ···················· | ···················· | ···················· |
| AB | 614 | hCoV-19/USA/NY-Wadsworth-288353-01/2020\|EPI_ISL_76 | ···················· | ···················· | ···················· | ···················· | ···················· | ···················· |
| AB | 615 | hCoV-19/USA/NY-Wadsworth-288367-01/2020\|EPI_ISL_76 | ···················· | ···················· | ···················· | ···················· | ···················· | ···················· |
| AB | 616 | hCoV-19/USA/NY-Wadsworth-288420-01/2020\|EPI_ISL_76 | ···················· | ···················· | ···················· | ···················· | ···················· | ···················· |
| AB | 617 | hCoV-19/USA/NY-Wadsworth-288425-01/2020\|EPI_ISL_76 | ···················· | ···················· | ···················· | ···················· | ···················· | ···················· |
| AB | 618 | hCoV-19/USA/NY-Wadsworth-288623-01/2020\|EPI_ISL_76 | ···················· | ···················· | ···················· | ···················· | ···················· | ···················· |
| AB | 619 | hCoV-19/USA/NY-Wadsworth-288783-01/2020\|EPI_ISL_76 | ···················· | ···················· | ···················· | ···················· | ···················· | ···················· |
| AB | 620 | hCoV-19/USA/NY-Wadsworth-288788-01/2020\|EPI_ISL_76 | ···················· | ···················· | ···················· | ···················· | ···················· | ··A················· |
| AB | 621 | hCoV-19/USA/NY-Wadsworth-288823-01/2020\|EPI_ISL_76 | ···················· | ···················· | ···················· | ···················· | ···················· | ···················· |
| AB | 622 | hCoV-19/USA/NY-Wadsworth-288824-01/2020\|EPI_ISL_76 | ···················· | ···················· | ···················· | ···················· | ···················· | ···················· |
| AB | 623 | hCoV-19/USA/NY-Wadsworth-288843-01/2020\|EPI_ISL_76 | ···················· | ···················· | ···················· | ···················· | ···················· | ···················· |
| AB | 624 | hCoV-19/USA/NY-Wadsworth-288854-01/2020\|EPI_ISL_76 | ···················· | ·A·················· | ···················· | ···················· | ···················· | ···················· |
| AB | 625 | hCoV-19/USA/NY-Wadsworth-288857-01/2020\|EPI_ISL_76 | ···················· | ···················· | ···················· | ···················· | ···················· | ···················· |
| AB | 626 | hCoV-19/USA/NY-Wadsworth-288888-01/2020\|EPI_ISL_76 | ···················· | ···················· | ···················· | ···················· | ···················· | ···················· |
| AB | 627 | hCoV-19/USA/NY-Wadsworth-288889-01/2020\|EPI_ISL_76 | ···················· | ···················· | ···················· | ···················· | ···················· | ···················· |
| AB | 628 | hCoV-19/USA/NY-Wadsworth-288890-01/2020\|EPI_ISL_76 | ···················· | ···················· | ···················· | ···················· | ···················· | ···················· |
| AB | 629 | hCoV-19/USA/NY-Wadsworth-288918-01/2020\|EPI_ISL_76 | ···················· | ·A·················· | ···················· | ···················· | ···················· | ···················· |
| AB | 630 | hCoV-19/USA/NY-Wadsworth-288935-01/2020\|EPI_ISL_76 | ···················· | ···················· | ···················· | ···················· | ···················· | ···················· |
| AB | 631 | hCoV-19/USA/NY-Wadsworth-288936-01/2020\|EPI_ISL_76 | ···················· | ···················· | ···················· | ···················· | ···················· | ···················· |

| | | | | | | | |
|---|---|---|---|---|---|---|---|
| AB | 632 hCoV-19/USA/NY-Wadsworth-289072-01/2020\|EPI_ISL_76 | ···················· | ···················· | ···················· | ···················· | ···················· | ···················· |
| AB | 633 hCoV-19/USA/NY-Wadsworth-289141-01/2020\|EPI_ISL_76 | ···················· | ···················· | ···················· | ···················· | ···················· | ···················· |
| AB | 634 hCoV-19/USA/NY-Wadsworth-289161-01/2020\|EPI_ISL_76 | ···················· | ···················· | ···················· | ···················· | ···················· | ···················· |
| AB | 635 hCoV-19/Japan/IC-0495/2020\|EPI_ISL_768704\|2020-12 | ···················· | ···················· | ···················· | ···················· | ···················· | ···················· |
| AB | 636 hCoV-19/Japan/IC-0496/2020\|EPI_ISL_768705\|2020-12 | ···················· | ···················· | ···················· | ···················· | ···················· | ···················· |
| AB | 637 hCoV-19/Japan/IC-0497/2020\|EPI_ISL_768706\|2020-12 | ···················· | ···················· | ···················· | ···················· | ···················· | ···················· |
| AB | 638 hCoV-19/Japan/IC-0500/2020\|EPI_ISL_768709\|2020-12 | ···················· | ···················· | ···················· | ···················· | ···················· | ···················· |
| AB | 639 hCoV-19/Japan/IC-0501/2020\|EPI_ISL_768710\|2020-12 | ···················· | ···················· | ···················· | ···················· | ···················· | ···················· |
| AB | 640 hCoV-19/Japan/IC-0502/2020\|EPI_ISL_768711\|2020-12 | ···················· | ···················· | ···················· | ···················· | ···················· | ···················· |
| AB | 641 hCoV-19/Japan/IC-0503/2020\|EPI_ISL_768712\|2020-12 | ···················· | ···················· | ···················· | ···················· | ···················· | ···················· |
| AB | 642 hCoV-19/Japan/IC-0504/2020\|EPI_ISL_768713\|2020-12 | ···················· | ···················· | ···················· | ···················· | ···················· | ···················· |
| AB | 643 hCoV-19/Japan/IC-0506/2020\|EPI_ISL_768715\|2020-12 | ···················· | ···················· | ···················· | ···················· | ···················· | ···················· |
| AB | 644 hCoV-19/Japan/IC-0507/2020\|EPI_ISL_768716\|2020-12 | ···················· | ·A·················· | ···················· | ···················· | ···················· | ···················· |
| AB | 645 hCoV-19/Japan/IC-0509/2020\|EPI_ISL_768718\|2020-12 | ···················· | ···················· | ···················· | ···················· | ···················· | ···················· |
| AB | 646 hCoV-19/Japan/IC-0510/2020\|EPI_ISL_768719\|2020-12 | ···················· | ···················· | ···················· | ···················· | ···················· | ···················· |
| AB | 647 hCoV-19/Japan/IC-0511/2020\|EPI_ISL_768720\|2020-12 | ···················· | ···················· | ···················· | ···················· | ···················· | ···················· |
| AB | 648 hCoV-19/Japan/IC-0512/2020\|EPI_ISL_768721\|2020-12 | ···················· | ···················· | ···················· | ···················· | ···················· | ···················· |
| AB | 649 hCoV-19/Japan/IC-0513/2020\|EPI_ISL_768722\|2020-12 | ···················· | ···················· | ···················· | ···················· | ···················· | ···················· |
| AB | 650 hCoV-19/Japan/IC-0514/2020\|EPI_ISL_768723\|2020-12 | ···················· | ···················· | ···················· | ···················· | ···················· | ···················· |
| AB | 651 hCoV-19/Japan/IC-0515/2020\|EPI_ISL_768724\|2020-12 | ···················· | ···················· | ···················· | ···················· | ···················· | ···················· |
| AB | 652 hCoV-19/Germany/TH-IIMK-CaSe-30/2020\|EPI_ISL_76872 | ···················· | ···················· | ···················· | ···················· | ···················· | ···················· |
| AB | 653 hCoV-19/Germany/TH-IIMK-CaSe-31/2020\|EPI_ISL_76872 | ···················· | ···················· | ···················· | ···················· | ···················· | ···················· |
| AB | 654 hCoV-19/Germany/TH-IIMK-CaSe-32/2020\|EPI_ISL_76872 | ···················· | ···················· | ···················· | ···················· | ···················· | ···················· |
| AB | 655 hCoV-19/Bangladesh/CHRF-0033/2020\|EPI_ISL_768729\|2 | ···················· | ···················· | ···················· | ···················· | ···················· | ···················· |
| AB | 656 hCoV-19/Bangladesh/CHRF-0036/2020\|EPI_ISL_768732\|2 | ···················· | ···················· | ···················· | ···················· | ···················· | ···················· |
| AB | 657 hCoV-19/Bangladesh/CHRF-0037/2020\|EPI_ISL_768733\|2 | ···················· | ···················· | ···················· | ···················· | ···················· | ···················· |
| AB | 658 hCoV-19/Bangladesh/CHRF-0041/2020\|EPI_ISL_768736\|2 | ···················· | ·A·················· | ···················· | ···················· | ···················· | ··A················· |
| AB | 659 hCoV-19/Bangladesh/CHRF-0065/2020\|EPI_ISL_768741\|2 | ···················· | ···················· | ···················· | ···················· | ···················· | ···················· |
| AB | 660 hCoV-19/Ireland/D-NVRL-MPJ90913/2020\|EPI_ISL_76874 | ···················· | ···················· | ···················· | ···················· | ···················· | ···················· |
| AB | 661 hCoV-19/Ireland/LH-NVRL-MPU16552/2020\|EPI_ISL_7687 | ···················· | ···················· | ···················· | ···················· | ···················· | ···················· |
| AB | 663 hCoV-19/Ireland/LH-NVRL-MPS62786/2020\|EPI_ISL_7687 | ···················· | ···················· | ···················· | ···················· | ···················· | ···················· |
| AB | 664 hCoV-19/Ireland/WH-NVRL-MPS15031/2020\|EPI_ISL_7687 | ···················· | ···················· | ···················· | ···················· | ···················· | ···················· |
| AB | 665 hCoV-19/Ireland/WH-NVRL-MPP60633/2020\|EPI_ISL_7687 | ···················· | ···················· | ···················· | ···················· | ···················· | ···················· |
| AB | 666 hCoV-19/Ireland/D-NVRL-AIIDV0086v1/2020\|EPI_ISL_76 | ···················· | ···················· | ···················· | ···················· | ···················· | ···················· |
| AB | 667 hCoV-19/Ireland/D-NVRL-AIIDV0432v2/2020\|EPI_ISL_76 | ···················· | ···················· | ···················· | ···················· | ···················· | ···················· |
| AB | 668 hCoV-19/Ireland/WW-NVRL-AIIDV0063v2/2020\|EPI_ISL_7 | ···················· | ···················· | ···················· | ···················· | ···················· | ···················· |
| AB | 669 hCoV-19/Ireland/D-NVRL-AIIDV0363v2/2020\|EPI_ISL_76 | ···················· | ···················· | ···················· | ···················· | ···················· | ···················· |
| AB | 670 hCoV-19/Ireland/D-NVRL-AIIDV0432v2-2/2020\|EPI_ISL_ | ···················· | ···················· | ···················· | ···················· | ···················· | ···················· |
| AB | 671 hCoV-19/Ireland/D-NVRL-AIIDV0380v1-2/2020\|EPI_ISL_ | ···················· | ···················· | ···················· | ···················· | ···················· | ···················· |
| AB | 672 hCoV-19/Ireland/D-NVRL-AIIDV1085v1/2020\|EPI_ISL_76 | ···················· | ···················· | ···················· | ···················· | ···················· | ···················· |
| AB | 673 hCoV-19/Ireland/D-NVRL-AIIDM0161v1/2020\|EPI_ISL_76 | ···················· | ···················· | ···················· | ···················· | ··T················· | ···················· |
| AB | 674 hCoV-19/Ireland/D-NVRL-AIIDV1088v1/2020\|EPI_ISL_76 | ···················· | ···················· | ···················· | ···················· | ···················· | ···················· |
| AB | 675 hCoV-19/Ireland/D-NVRL-AIIDM0156v1/2020\|EPI_ISL_76 | ···················· | ···················· | ···················· | ···················· | ··T················· | ···················· |
| AB | 676 hCoV-19/Ireland/D-NVRL-AIIDVNA883v1/2020\|EPI_ISL_7 | ···················· | ···················· | ···················· | ···················· | ···················· | ···················· |
| AB | 677 hCoV-19/Ireland/D-NVRL-AIIDM0162v1/2020\|EPI_ISL_76 | ···················· | ···················· | ···················· | ···················· | ···················· | ···················· |
| AB | 678 hCoV-19/Ireland/WW-NVRL-AIIDV1098v1/2020\|EPI_ISL_7 | ···················· | ···················· | ···················· | ···················· | ···················· | ···················· |
| AB | 679 hCoV-19/Ireland/WW-NVRL-AIIDV1099v1/2020\|EPI_ISL_7 | ···················· | ···················· | ···················· | ···················· | ···················· | ···················· |
| AB | 680 hCoV-19/Ireland/WW-NVRL-AIIDV1206v1/2020\|EPI_ISL_7 | ···················· | ···················· | ···················· | ···················· | ···················· | ···················· |
| AB | 681 hCoV-19/Ireland/D-NVRL-AIIDV1190v1/2020\|EPI_ISL_76 | ···················· | ···················· | ···················· | ···················· | ···················· | ···················· |
| AB | 682 hCoV-19/Ireland/D-NVRL-AIIDV1322v1/2020\|EPI_ISL_76 | ···················· | ···················· | ···················· | ···················· | ···················· | ···················· |
| AB | 683 hCoV-19/Ireland/D-NVRL-AIIDM0167v1/2020\|EPI_ISL_76 | ···················· | ···················· | ···················· | ···················· | ···················· | ···················· |
| AB | 684 hCoV-19/Ireland/D-NVRL-AIIDV1213v1/2020\|EPI_ISL_76 | ···················· | ···················· | ···················· | ···················· | ···················· | ···················· |
| AB | 685 hCoV-19/Ireland/D-NVRL-AIIDV1324v1/2020\|EPI_ISL_76 | ···················· | ···················· | ···················· | ···················· | ···················· | ···················· |
| AB | 686 hCoV-19/Ireland/D-NVRL-AIIDV1325v1/2020\|EPI_ISL_76 | ···················· | ···················· | ···················· | ···················· | ···················· | ···················· |
| AB | 687 hCoV-19/Ireland/D-NVRL-AIIDV1239v1/2020\|EPI_ISL_76 | ···················· | ···················· | ···················· | ···················· | ···················· | ···················· |
| AB | 688 hCoV-19/Ireland/D-NVRL-AIIDV1252v1/2020\|EPI_ISL_76 | ···················· | ···················· | ···················· | ···················· | ···················· | ···················· |
| AB | 689 hCoV-19/Ireland/WW-NVRL-AIIDV1257v1/2020\|EPI_ISL_7 | ···················· | ···················· | ···················· | ···················· | ···················· | ···················· |
| AB | 690 hCoV-19/Ireland/D-NVRL-AIIDM0175v1/2020\|EPI_ISL_76 | ···················· | ···················· | ···················· | ···················· | ···················· | ···················· |
| AB | 691 hCoV-19/Ireland/D-NVRL-AIIDV1333v1/2020\|EPI_ISL_76 | ···················· | ···················· | ···················· | ···················· | ···················· | ···················· |
| AB | 692 hCoV-19/Ireland/D-NVRL-AIIDV1332v1/2020\|EPI_ISL_76 | ···················· | ···················· | ···················· | ···················· | ···················· | ···················· |
| AB | 693 hCoV-19/Ireland/D-NVRL-AIIDV1268v1/2020\|EPI_ISL_76 | ···················· | ···················· | ···················· | ···················· | ···················· | ···················· |
| AB | 694 hCoV-19/Ireland/D-NVRL-AIIDM0179v1/2020\|EPI_ISL_76 | ···················· | ···················· | ···················· | ···················· | ···················· | ···················· |
| AB | 695 hCoV-19/Ireland/D-NVRL-AIIDM0180v1/2020\|EPI_ISL_76 | ···················· | ···················· | ···················· | ···················· | ···················· | ···················· |
| AB | 696 hCoV-19/Ireland/D-NVRL-AIIDV1335v1/2020\|EPI_ISL_76 | ···················· | ···················· | ···················· | ···················· | ···················· | ···················· |
| AB | 697 hCoV-19/Ireland/WW-NVRL-AIIDV1272v1/2020\|EPI_ISL_7 | ···················· | ···················· | ···················· | ···················· | ···················· | ···················· |
| AB | 698 hCoV-19/Ireland/D-NVRL-AIIDM0184v1/2020\|EPI_ISL_76 | ···················· | ···················· | ···················· | ···················· | ···················· | ···················· |
| AB | 699 hCoV-19/Ireland/D-NVRL-AIIDV1278v1/2020\|EPI_ISL_76 | ···················· | ···················· | ···················· | ···················· | ···················· | ···················· |
| AB | 700 hCoV-19/Ireland/WW-NVRL-AIIDV1296v1/2020\|EPI_ISL_7 | ···················· | ···················· | ···················· | ···················· | ···················· | ···················· |
| AB | 701 hCoV-19/Ireland/WW-NVRL-AIIDV1295v1/2020\|EPI_ISL_7 | ···················· | ···················· | ···················· | ···················· | ···················· | ···················· |
| AB | 702 hCoV-19/Ireland/WX-NVRL-AIIDW001v1/2020\|EPI_ISL_76 | ···················· | ···················· | ···················· | ···················· | ···················· | ···················· |
| AB | 703 hCoV-19/Ireland/D-NVRL-AIIDV1340v1/2020\|EPI_ISL_76 | ···················· | ···················· | ···················· | ···················· | ···················· | ···················· |
| AB | 704 hCoV-19/Ireland/D-NVRL-AIIDV1339v1/2020\|EPI_ISL_76 | ···················· | ···················· | ···················· | ···················· | ···················· | ···················· |
| AB | 705 hCoV-19/Germany/HH-hpi-p1528/2020\|EPI_ISL_775905\|2 | ···················· | ···················· | ···················· | ···················· | ···················· | ···················· |
| AB | 706 hCoV-19/Germany/HH-hpi-p1513/2020\|EPI_ISL_775906\|2 | ···················· | ···················· | ···················· | ···················· | ···················· | ···················· |
| AB | 707 hCoV-19/Germany/HH-hpi-p1432/2020\|EPI_ISL_775953\|2 | ···················· | ···················· | ···················· | ···················· | ···················· | ···················· |
| AB | 708 hCoV-19/Germany/HH-hpi-p1434/2020\|EPI_ISL_775954\|2 | ···················· | ···················· | ···················· | ···················· | ···················· | ···················· |
| AB | 709 hCoV-19/Germany/HH-hpi-p1435/2020\|EPI_ISL_775955\|2 | ···················· | ···················· | ···················· | ···················· | ···················· | ···················· |
| AB | 710 hCoV-19/Germany/HH-hpi-p1436/2020\|EPI_ISL_775956\|2 | ···················· | ···················· | ···················· | ···················· | ···················· | ···················· |
| AB | 711 hCoV-19/Germany/HH-hpi-p1437/2020\|EPI_ISL_775957\|2 | ···················· | ···················· | ···················· | ···················· | ···················· | ···················· |
| AB | 712 hCoV-19/Germany/HH-hpi-p1438/2020\|EPI_ISL_775958\|2 | ···················· | ···················· | ···················· | ···················· | ···················· | ···················· |

| | | | | | | | |
|---|---|---|---|---|---|---|---|
| AB | 713 hCoV-19/Germany/HH-hpi-p1439/2020\|EPI_ISL_775959\|2 | ···················· | ···················· | ···················· | ···················· | ···················· | ···················· |
| AB | 714 hCoV-19/Germany/HH-hpi-p1441/2020\|EPI_ISL_775960\|2 | ···················· | ···················· | ···················· | ···················· | ···················· | ···················· |
| AB | 715 hCoV-19/Germany/HH-hpi-p1442/2020\|EPI_ISL_775961\|2 | ···················· | ···················· | ···················· | ···················· | ···················· | ···················· |
| AB | 716 hCoV-19/Germany/HH-hpi-p1443/2020\|EPI_ISL_775962\|2 | ···················· | ···················· | ···················· | ···················· | ···················· | ···················· |
| AB | 717 hCoV-19/Germany/HH-hpi-p1444/2020\|EPI_ISL_775963\|2 | ···················· | ···················· | ···················· | ···················· | ···················· | ···················· |
| AB | 718 hCoV-19/Germany/HH-hpi-p1445/2020\|EPI_ISL_775964\|2 | ···················· | ···················· | ···················· | ···················· | ···················· | ···················· |
| AB | 719 hCoV-19/Germany/HH-hpi-p1446/2020\|EPI_ISL_775965\|2 | ···················· | ···················· | ···················· | ···················· | ···················· | ···················· |
| AB | 720 hCoV-19/Germany/HH-hpi-p1447/2020\|EPI_ISL_775966\|2 | ···················· | ···················· | ···················· | ···················· | ···················· | ···················· |
| AB | 721 hCoV-19/Germany/HH-hpi-p1448/2020\|EPI_ISL_775967\|2 | ···················· | ···················· | ···················· | ···················· | ···················· | ···················· |
| AB | 722 hCoV-19/Germany/HH-hpi-p1449/2020\|EPI_ISL_775968\|2 | ···················· | ···················· | ···················· | ···················· | ···················· | ···················· |
| AB | 723 hCoV-19/Germany/HH-hpi-p1452/2020\|EPI_ISL_775969\|2 | ···················· | ···················· | ···················· | ···················· | ···················· | ···················· |
| AB | 724 hCoV-19/Germany/HH-hpi-p1455/2020\|EPI_ISL_775970\|2 | ···················· | ···················· | ···················· | ···················· | ···················· | ···················· |
| AB | 725 hCoV-19/Germany/HH-hpi-p1456/2020\|EPI_ISL_775971\|2 | ···················· | ···················· | ···················· | ···················· | ···················· | ···················· |
| AB | 726 hCoV-19/Germany/HH-hpi-p1457/2020\|EPI_ISL_775972\|2 | ···················· | ···················· | ···················· | ···················· | ···················· | ···················· |
| AB | 727 hCoV-19/Germany/HH-hpi-p1460/2020\|EPI_ISL_775973\|2 | ···················· | ···················· | ···················· | ···················· | ···················· | ···················· |
| AB | 728 hCoV-19/Germany/HH-hpi-p1461/2020\|EPI_ISL_775974\|2 | ···················· | ···················· | ···················· | ···················· | ···················· | ···················· |
| AB | 729 hCoV-19/Germany/HH-hpi-p1462/2020\|EPI_ISL_775975\|2 | ···················· | ···················· | ···················· | ···················· | ···················· | ···················· |
| AB | 730 hCoV-19/Germany/HH-hpi-p1463/2020\|EPI_ISL_775976\|2 | ···················· | ···················· | ···················· | ···················· | ···················· | ···················· |
| AB | 732 hCoV-19/Germany/HH-hpi-p1467/2020\|EPI_ISL_775979\|2 | ···················· | ···················· | ···················· | ···················· | ···················· | ···················· |
| AB | 733 hCoV-19/Germany/HH-hpi-p1475/2020\|EPI_ISL_775986\|2 | ···················· | ···················· | ···················· | ···················· | ···················· | ···················· |
| AB | 734 hCoV-19/Germany/HH-hpi-p1476/2020\|EPI_ISL_775987\|2 | ···················· | ···················· | ···················· | ···················· | ···················· | ···················· |
| AB | 735 hCoV-19/Germany/HH-hpi-p1477/2020\|EPI_ISL_775988\|2 | ···················· | ···················· | ···················· | ···················· | ···················· | ···················· |
| AB | 736 hCoV-19/Germany/HH-hpi-p1478/2020\|EPI_ISL_775989\|2 | ···················· | ···················· | ···················· | ···················· | ···················· | ···················· |
| AB | 737 hCoV-19/Germany/HH-hpi-p1479/2020\|EPI_ISL_775990\|2 | ···················· | ···················· | ···················· | ···················· | ···················· | ···················· |
| AB | 738 hCoV-19/Germany/HH-hpi-p1480/2020\|EPI_ISL_775991\|2 | ···················· | ···················· | ···················· | ···················· | ···················· | ···················· |
| AB | 739 hCoV-19/Germany/HH-hpi-p1481/2020\|EPI_ISL_775992\|2 | ···················· | ···················· | ···················· | ···················· | ···················· | ···················· |
| AB | 740 hCoV-19/Germany/HH-hpi-p1483/2020\|EPI_ISL_775993\|2 | ···················· | ···················· | ···················· | ···················· | ···················· | ···················· |
| AB | 741 hCoV-19/Germany/HH-hpi-p1484/2020\|EPI_ISL_775994\|2 | ···················· | ···················· | ···················· | ···················· | ···················· | ···················· |
| AB | 742 hCoV-19/Germany/HH-hpi-p1485/2020\|EPI_ISL_775995\|2 | ···················· | ···················· | ···················· | ···················· | ···················· | ···················· |
| AB | 743 hCoV-19/Germany/HH-hpi-p1486/2020\|EPI_ISL_775996\|2 | ···················· | ···················· | ···················· | ···················· | ···················· | ···················· |
| AB | 744 hCoV-19/England/CAMC-CB6FB5/2020\|EPI_ISL_778100\|20 | ···················· | ···················· | ···················· | ···················· | ···················· | ···················· |
| AB | 745 hCoV-19/England/QEUH-CC2E04/2020\|EPI_ISL_778102\|20 | ···················· | ···················· | ···················· | ···················· | ···················· | ···················· |
| AB | 746 hCoV-19/England/ALDP-CC07DE/2020\|EPI_ISL_778104\|20 | ···················· | ···················· | ···················· | ···················· | ···················· | ···················· |
| AB | 747 hCoV-19/England/QEUH-CC28AC/2020\|EPI_ISL_778111\|20 | ···················· | ···················· | ···················· | ···················· | ···················· | ···················· |
| AB | 748 hCoV-19/England/QEUH-CC25E1/2020\|EPI_ISL_778117\|20 | ···················· | ···················· | ···················· | ···················· | ······T············· | ···················· |
| AB | 749 hCoV-19/England/QEUH-CC28BB/2020\|EPI_ISL_778119\|20 | ···················· | ···················· | ···················· | ···················· | ···················· | ···················· |
| AB | 750 hCoV-19/England/QEUH-CC2D8F/2020\|EPI_ISL_778122\|20 | ···················· | ···················· | ···················· | ···················· | ···················· | ···················· |
| AB | 751 hCoV-19/England/CAMC-CB8E5C/2020\|EPI_ISL_778126\|20 | ···················· | ···················· | ···················· | ···················· | ···················· | ···················· |
| AB | 752 hCoV-19/England/QEUH-CC2DDA/2020\|EPI_ISL_778129\|20 | ···················· | ···················· | ···················· | ···················· | ···················· | ···················· |
| AB | 753 hCoV-19/England/ALDP-CC0738/2020\|EPI_ISL_778132\|20 | ···················· | ···················· | ···················· | ···················· | ···················· | ···················· |
| AB | 754 hCoV-19/England/QEUH-CC3067/2020\|EPI_ISL_778133\|20 | ···················· | ···················· | ···················· | ···················· | ···················· | ···················· |
| AB | 755 hCoV-19/England/CAMC-CB8B82/2020\|EPI_ISL_778134\|20 | ···················· | ···················· | ···················· | ···················· | ······T············· | ···················· |
| AB | 756 hCoV-19/England/QEUH-CC2D52/2020\|EPI_ISL_778149\|20 | ···················· | ···················· | ···················· | ···················· | ···················· | ···················· |
| AB | 757 hCoV-19/England/QEUH-CC2402/2020\|EPI_ISL_778153\|20 | ···················· | ···················· | ···················· | ···················· | ···················· | ···················· |
| AB | 758 hCoV-19/England/QEUH-CC2D34/2020\|EPI_ISL_778154\|20 | ···················· | ···················· | ···················· | ···················· | ···················· | ···················· |
| AB | 759 hCoV-19/England/QEUH-CC2DBC/2020\|EPI_ISL_778156\|20 | ···················· | ···················· | ···················· | ···················· | ···················· | ···················· |
| AB | 760 hCoV-19/England/QEUH-CC2E40/2020\|EPI_ISL_778159\|20 | ···················· | ···················· | ···················· | ···················· | ···················· | ···················· |
| AB | 761 hCoV-19/England/ALDP-CC099C/2020\|EPI_ISL_778164\|20 | ···················· | ···················· | ···················· | ···················· | ···················· | ···················· |
| AB | 762 hCoV-19/England/ALDP-CBF22C/2020\|EPI_ISL_778166\|20 | ···················· | ···················· | ··········T········· | ···················· | ···················· | ···················· |
| AB | 763 hCoV-19/England/QEUH-CC22AE/2020\|EPI_ISL_778167\|20 | ···················· | ·········T·········· | ···················· | ···················· | ···················· | ···················· |
| AB | 764 hCoV-19/England/QEUH-CC2851/2020\|EPI_ISL_778168\|20 | ···················· | ···················· | ···················· | ···················· | ···················· | ···················· |
| AB | 765 hCoV-19/England/CAMC-CB8EF2/2020\|EPI_ISL_778174\|20 | ···················· | ···················· | ···················· | ···················· | ······T············· | ···················· |
| AB | 766 hCoV-19/England/QEUH-CC2D16/2020\|EPI_ISL_778176\|20 | ···················· | ···················· | ···················· | ···················· | ···················· | ···················· |
| AB | 767 hCoV-19/England/CAMC-CB8B91/2020\|EPI_ISL_778179\|20 | ···················· | ···················· | ···················· | ···················· | ···················· | ···················· |
| AB | 768 hCoV-19/England/ALDP-CC07A1/2020\|EPI_ISL_778183\|20 | ···················· | ···················· | ···················· | ···················· | ···················· | ···················· |
| AB | 769 hCoV-19/South Korea/KDCA0249/2020\|EPI_ISL_760014\|2 | ···················· | ···················· | ···················· | ···················· | ···················· | ···················· |
| AB | 770 hCoV-19/South Korea/KDCA0250/2020\|EPI_ISL_760015\|2 | ···················· | ···················· | ···················· | ···················· | ···················· | ···················· |
| AB | 771 hCoV-19/South Korea/KDCA0251/2020\|EPI_ISL_760016\|2 | ···················· | ···················· | ···················· | ···················· | ···················· | ···················· |
| AB | 772 hCoV-19/South Korea/KDCA0252/2020\|EPI_ISL_760017\|2 | ···················· | ···················· | ···················· | ···················· | ···················· | ···················· |
| AB | 773 hCoV-19/South Korea/KDCA0253/2020\|EPI_ISL_760018\|2 | ···················· | ···················· | ···················· | ···················· | ···················· | ···················· |
| AB | 774 hCoV-19/South Korea/KDCA0254/2020\|EPI_ISL_760019\|2 | ···················· | ···················· | ···················· | ···················· | ···················· | ···················· |
| AB | 775 hCoV-19/South Korea/KDCA0255/2020\|EPI_ISL_760020\|2 | ···················· | ···················· | ···················· | ···················· | ···················· | ···················· |
| AB | 776 hCoV-19/South Korea/KDCA0256/2020\|EPI_ISL_760021\|2 | ···················· | ···················· | ···················· | ···················· | ···················· | ···················· |
| AB | 777 hCoV-19/South Korea/KDCA0257/2020\|EPI_ISL_760022\|2 | ···················· | ···················· | ···················· | ···················· | ···················· | ···················· |
| AB | 778 hCoV-19/South Korea/KDCA0258/2020\|EPI_ISL_760023\|2 | ···················· | ···················· | ···················· | ···················· | ···················· | ···················· |
| AB | 779 hCoV-19/South Korea/KDCA0259/2020\|EPI_ISL_760024\|2 | ···················· | ···················· | ···················· | ···················· | ···················· | ···················· |
| AB | 780 hCoV-19/South Korea/KDCA0260/2020\|EPI_ISL_760025\|2 | ···················· | ···················· | ···················· | ···················· | ···················· | ···················· |
| AB | 781 hCoV-19/South Korea/KDCA0261/2020\|EPI_ISL_760026\|2 | ···················· | ···················· | ···················· | ···················· | ···················· | ···················· |
| AB | 782 hCoV-19/South Korea/KDCA0262/2020\|EPI_ISL_760027\|2 | ···················· | ···················· | ···················· | ···················· | ···················· | ···················· |
| AB | 783 hCoV-19/South Korea/KDCA0263/2020\|EPI_ISL_760028\|2 | ···················· | ···················· | ···················· | ···················· | ···················· | ···················· |
| AB | 784 hCoV-19/South Korea/KDCA0264/2020\|EPI_ISL_760029\|2 | ···················· | ···················· | ···················· | ···················· | ···················· | ···················· |
| AB | 785 hCoV-19/South Korea/KDCA0265/2020\|EPI_ISL_760030\|2 | ···················· | ···················· | ···················· | ···················· | ···················· | ···················· |
| AB | 786 hCoV-19/Hong Kong/VM20109237/2020\|EPI_ISL_760031\|2 | ···················· | ···················· | ···················· | ···················· | ···················· | ···················· |
| AB | 787 hCoV-19/Hong Kong/VM20123554/2020\|EPI_ISL_760032\|2 | ···················· | ·A·················· | ···················· | ···················· | ···················· | ···A················ |
| AB | 788 hCoV-19/Hong Kong/VM20123555/2020\|EPI_ISL_760033\|2 | ···················· | ·A·················· | ···················· | ···················· | ···················· | ···A················ |
| AB | 789 hCoV-19/Hong Kong/VM20123558/2020\|EPI_ISL_760034\|2 | ···················· | ·A·················· | ···················· | ···················· | ···················· | ···A················ |
| AB | 790 hCoV-19/Hong Kong/VM20123559/2020\|EPI_ISL_760035\|2 | ···················· | ·A·················· | ···················· | ···················· | ···················· | ···A················ |
| AB | 791 hCoV-19/Hong Kong/VM20123560/2020\|EPI_ISL_760036\|2 | ···················· | ·A·················· | ···················· | ···················· | ···················· | ···A················ |
| AB | 792 hCoV-19/Hong Kong/VM20126869/2020\|EPI_ISL_760037\|2 | ···················· | ·A·················· | ···················· | ···················· | ···················· | ···A················ |
| AB | 793 hCoV-19/Hong Kong/VM20127499/2020\|EPI_ISL_760038\|2 | ···················· | ·A·················· | ···················· | ···················· | ···················· | ···A················ |

| | | | | | | | |
|---|---|---|---|---|---|---|---|
| AB | 794 hCoV-19/Hong Kong/VM20127572/2020\|EPI_ISL_760039\|2 | ···················· | ·A·················· | ···················· | ···················· | ···················· | ··A················· |
| AB | 795 hCoV-19/Hong Kong/VM20127573/2020\|EPI_ISL_760040\|2 | ···················· | ·A·················· | ···················· | ···················· | ···················· | ··A················· |
| AB | 796 hCoV-19/Hong Kong/VM20127769/2020\|EPI_ISL_760041\|2 | ···················· | ·A·················· | ···················· | ···················· | ···················· | ··A················· |
| AB | 797 hCoV-19/Hong Kong/VM20127935/2020\|EPI_ISL_760042\|2 | ···················· | ·A·················· | ···················· | ···················· | ···················· | ··A················· |
| AB | 798 hCoV-19/Hong Kong/VM20127956/2020\|EPI_ISL_760043\|2 | ···················· | ·A·················· | ···················· | ···················· | ···················· | ··A················· |
| AB | 799 hCoV-19/Hong Kong/VM20128243/2020\|EPI_ISL_760044\|2 | ···················· | ·A·················· | ···················· | ···················· | ···················· | ··A················· |
| AB | 800 hCoV-19/Hong Kong/VB20209003/2020\|EPI_ISL_760045\|2 | ···················· | ·A·················· | ···················· | ···················· | ···················· | ··A················· |
| AB | 801 hCoV-19/Hong Kong/VB20209026/2020\|EPI_ISL_760046\|2 | ···················· | ·A·················· | ···················· | ···················· | ···················· | ··A················· |
| AB | 802 hCoV-19/Hong Kong/VM20128899/2020\|EPI_ISL_760047\|2 | ···················· | ·A·················· | ···················· | ···················· | ···················· | ··A················· |
| AB | 803 hCoV-19/Hong Kong/VM20129245/2020\|EPI_ISL_760048\|2 | ···················· | ·A·················· | ···················· | ···················· | ···················· | ··A················· |
| AB | 804 hCoV-19/Hong Kong/VM20129415/2020\|EPI_ISL_760049\|2 | ···················· | ·A·················· | ···················· | ···················· | ···················· | ··A················· |
| AB | 805 hCoV-19/Hong Kong/VB20211407/2020\|EPI_ISL_760050\|2 | ···················· | ·A·················· | ···················· | ···················· | ···················· | ··A················· |
| AB | 806 hCoV-19/Hong Kong/VB20211660/2020\|EPI_ISL_760051\|2 | ···················· | ·A·················· | ···················· | ···················· | ···················· | ··A················· |
| AB | 807 hCoV-19/Hong Kong/VB20211661/2020\|EPI_ISL_760052\|2 | ···················· | ·A·················· | ···················· | ···················· | ···················· | ··A················· |
| AB | 808 hCoV-19/Hong Kong/VM20130028/2020\|EPI_ISL_760053\|2 | ···················· | ·A·················· | ···················· | ···················· | ···················· | ··A················· |
| AB | 809 hCoV-19/Hong Kong/VM20130432/2020\|EPI_ISL_760054\|2 | ···················· | ·A·················· | ···················· | ···················· | ···················· | ··A················· |
| AB | 810 hCoV-19/Hong Kong/VM20131073/2020\|EPI_ISL_760055\|2 | ···················· | ·A·················· | ···················· | ···················· | ···················· | ··A················· |
| AB | 811 hCoV-19/Hong Kong/VB20214929/2020\|EPI_ISL_760056\|2 | ···················· | ·A·················· | ···················· | ···················· | ···················· | ··A················· |
| AB | 812 hCoV-19/Hong Kong/VB20219976/2020\|EPI_ISL_760057\|2 | ···················· | ·A·················· | ···················· | ···················· | ···················· | ··A················· |
| AB | 813 hCoV-19/Hong Kong/VB20220907/2020\|EPI_ISL_760058\|2 | ···················· | ·A·················· | ···················· | ···················· | ···················· | ··A················· |
| AB | 814 hCoV-19/South Korea/KDCA0266/2020\|EPI_ISL_760060\|2 | ···················· | ···················· | ···················· | ···················· | ···················· | ···················· |
| AB | 815 hCoV-19/South Korea/KDCA0267/2020\|EPI_ISL_760061\|2 | ···················· | ···················· | ···················· | ···················· | ···················· | ···················· |
| AB | 816 hCoV-19/South Korea/KDCA0268/2020\|EPI_ISL_760062\|2 | ···················· | ···················· | ···················· | ···················· | ···················· | ···················· |
| AB | 817 hCoV-19/South Korea/KDCA0269/2020\|EPI_ISL_760063\|2 | ···················· | ·············A······ | ···················· | ···················· | ···················· | ···················· |
| AB | 818 hCoV-19/South Korea/KDCA0270/2020\|EPI_ISL_760064\|2 | ···················· | ·········A·········· | ···················· | ···················· | ···················· | ···················· |
| AB | 819 hCoV-19/South Korea/KDCA0271/2020\|EPI_ISL_760065\|2 | ···················· | ···················· | ···················· | ···················· | ···················· | ···················· |
| AB | 820 hCoV-19/South Korea/KDCA0272/2020\|EPI_ISL_760066\|2 | ···················· | ·············A······ | ···················· | ···················· | ···················· | ···················· |
| AB | 821 hCoV-19/South Korea/KDCA0273/2020\|EPI_ISL_760067\|2 | ···················· | ···················· | ···················· | ···················· | ···················· | ···················· |
| AB | 822 hCoV-19/South Korea/KDCA0274/2020\|EPI_ISL_760068\|2 | ···················· | ···················· | ···················· | ···················· | ···················· | ···················· |
| AB | 823 hCoV-19/South Korea/KDCA0275/2020\|EPI_ISL_760069\|2 | ···················· | ···················· | ···················· | ···················· | ···················· | ···················· |
| AB | 824 hCoV-19/South Korea/KDCA0276/2020\|EPI_ISL_760070\|2 | ···················· | ···················· | ···················· | ···················· | ···················· | ···················· |
| AB | 825 hCoV-19/South Korea/KDCA0277/2020\|EPI_ISL_760071\|2 | ···················· | ···················· | ···················· | ···················· | ···················· | ···················· |
| AB | 826 hCoV-19/South Korea/KDCA0278/2020\|EPI_ISL_760072\|2 | ···················· | ···················· | ···················· | ···················· | ···················· | ···················· |
| AB | 827 hCoV-19/South Korea/KDCA0280/2020\|EPI_ISL_760074\|2 | ···················· | ···················· | ···················· | ···················· | ···················· | ···················· |
| AB | 828 hCoV-19/South Korea/KDCA0281/2020\|EPI_ISL_760075\|2 | ···················· | ···················· | ···················· | ···················· | ···················· | ···················· |
| AB | 829 hCoV-19/South Korea/KDCA0282/2020\|EPI_ISL_760076\|2 | ···················· | ···················· | ···················· | ···················· | ···················· | ···················· |
| AB | 830 hCoV-19/South Korea/KDCA0283/2020\|EPI_ISL_760077\|2 | ···················· | ···················· | ···················· | ···················· | ······T············· | ···················· |
| AB | 831 hCoV-19/South Korea/KDCA0284/2020\|EPI_ISL_760078\|2 | ···················· | ···················· | ···················· | ···················· | ···················· | ···················· |
| AB | 832 hCoV-19/South Korea/KDCA0285/2020\|EPI_ISL_760079\|2 | ···················· | ···················· | ···················· | ···················· | ······T············· | ···················· |
| AB | 833 hCoV-19/South Korea/KDCA0286/2020\|EPI_ISL_760080\|2 | ···················· | ···················· | ···················· | ···················· | ······T············· | ···················· |
| AB | 834 hCoV-19/South Korea/KDCA0287/2020\|EPI_ISL_760081\|2 | ···················· | ···················· | ···················· | ···················· | ···················· | ···················· |
| AB | 835 hCoV-19/South Korea/KDCA0288/2020\|EPI_ISL_760082\|2 | ···················· | ···················· | ···················· | ···················· | ···················· | ···················· |
| AB | 836 hCoV-19/South Korea/KDCA0289/2020\|EPI_ISL_760083\|2 | ···················· | ···················· | ···················· | ···················· | ···················· | ···················· |
| AB | 837 hCoV-19/South Korea/KDCA0290/2020\|EPI_ISL_760084\|2 | ···················· | ···················· | ···················· | ···················· | ···················· | ···················· |
| AB | 838 hCoV-19/South Korea/KDCA0291/2020\|EPI_ISL_760085\|2 | ···················· | ···················· | ···················· | ···················· | ···················· | ···················· |
| AB | 839 hCoV-19/South Korea/KDCA0292/2020\|EPI_ISL_760086\|2 | ···················· | ···················· | ···················· | ···················· | ···················· | ···················· |
| AB | 840 hCoV-19/South Korea/KDCA0293/2020\|EPI_ISL_760087\|2 | ···················· | ···················· | ···················· | ···················· | ···················· | ···················· |
| AB | 841 hCoV-19/South Korea/KDCA0294/2020\|EPI_ISL_760088\|2 | ···················· | ···················· | ···················· | ···················· | ···················· | ···················· |
| AB | 842 hCoV-19/South Korea/KDCA0295/2020\|EPI_ISL_760089\|2 | ···················· | ···················· | ···················· | ···················· | ···················· | ···················· |
| AB | 843 hCoV-19/South Korea/KDCA0296/2020\|EPI_ISL_760090\|2 | ···················· | ···················· | ···················· | ···················· | ···················· | ···················· |
| AB | 844 hCoV-19/South Korea/KDCA0297/2020\|EPI_ISL_760091\|2 | ···················· | ···················· | ···················· | ···················· | ···················· | ···················· |
| AB | 845 hCoV-19/South Korea/KDCA0298/2020\|EPI_ISL_760092\|2 | ···················· | ···················· | ···················· | ···················· | ···················· | ···················· |
| AB | 846 hCoV-19/South Korea/KDCA0299/2020\|EPI_ISL_760093\|2 | ···················· | ···················· | ···················· | ···················· | ···················· | ···················· |
| AB | 847 hCoV-19/South Korea/KDCA0300/2020\|EPI_ISL_760094\|2 | ···················· | ···················· | ···················· | ···················· | ···················· | ···················· |
| AB | 848 hCoV-19/South Korea/KDCA0301/2020\|EPI_ISL_760095\|2 | ···················· | ···················· | ···················· | ···················· | ···················· | ···················· |
| AB | 849 hCoV-19/South Korea/KDCA0310/2020\|EPI_ISL_760096\|2 | ···················· | ···················· | ···················· | ···················· | ···················· | ···················· |
| AB | 850 hCoV-19/South Korea/KDCA0311/2020\|EPI_ISL_760097\|2 | ···················· | ···················· | ···················· | ···················· | ···················· | ···················· |
| AB | 851 hCoV-19/South Korea/KDCA0312/2020\|EPI_ISL_760098\|2 | ···················· | ···················· | ···················· | ···················· | ···················· | ···················· |
| AB | 852 hCoV-19/South Korea/KDCA0313/2020\|EPI_ISL_760099\|2 | ···················· | ···················· | ···················· | ···················· | ···················· | ···················· |
| AB | 853 hCoV-19/England/ALDP-CB5C82/2020\|EPI_ISL_761300\|20 | ···················· | ···················· | ···················· | ···················· | ···················· | ···················· |
| AB | 854 hCoV-19/England/ALDP-CB3659/2020\|EPI_ISL_761301\|20 | ···················· | ···················· | ···················· | ···················· | ···················· | ···················· |
| AB | 855 hCoV-19/England/QEUH-CADF0C/2020\|EPI_ISL_761302\|20 | ···················· | ···················· | ···················· | ···················· | ···················· | ···················· |
| AB | 856 hCoV-19/England/ALDP-CB4A3E/2020\|EPI_ISL_761306\|20 | ···················· | ···················· | ···················· | ···················· | ···················· | ···················· |
| AB | 857 hCoV-19/England/ALDP-CB67AE/2020\|EPI_ISL_761309\|20 | ···················· | ···················· | ···················· | ···················· | ···················· | ···················· |
| AB | 858 hCoV-19/England/ALDP-CB4A01/2020\|EPI_ISL_761310\|20 | ···················· | ·A·················· | ···················· | ···················· | ···················· | ··A················· |
| AB | 859 hCoV-19/England/ALDP-CB412A/2020\|EPI_ISL_761311\|20 | ···················· | ···················· | ···················· | ···················· | ···················· | ···················· |
| AB | 860 hCoV-19/England/ALDP-CB3DFA/2020\|EPI_ISL_761345\|20 | ···················· | ···················· | ···················· | ···················· | ···················· | ···················· |
| AB | 861 hCoV-19/England/ALDP-CB3C57/2020\|EPI_ISL_761346\|20 | ···················· | ···················· | ···················· | ···················· | ···················· | ···················· |
| AB | 862 hCoV-19/England/ALDP-CB51ED/2020\|EPI_ISL_761347\|20 | ···················· | ···················· | ···················· | ···················· | ···················· | ···················· |
| AB | 863 hCoV-19/England/QEUH-CAD986/2020\|EPI_ISL_761348\|20 | ···················· | ···················· | ···················· | ···················· | ···················· | ···················· |
| AB | 864 hCoV-19/England/ALDP-CB4913/2020\|EPI_ISL_761349\|20 | ···················· | ···················· | ···················· | ···················· | ···················· | ···················· |
| AB | 865 hCoV-19/England/QEUH-CAD7AA/2020\|EPI_ISL_761351\|20 | ···················· | ···················· | ···················· | ···················· | ···················· | ···················· |
| AB | 866 hCoV-19/England/ALDP-CB402D/2020\|EPI_ISL_761352\|20 | ···················· | ···················· | ···················· | ···················· | ···················· | ···················· |
| AB | 867 hCoV-19/England/QEUH-CADC6F/2020\|EPI_ISL_761353\|20 | ···················· | ···················· | ···················· | ···················· | ···················· | ···················· |
| AB | 868 hCoV-19/England/QEUH-CADDB7/2020\|EPI_ISL_761355\|20 | ···················· | ···················· | ···················· | ···················· | ······T············· | ···················· |
| AB | 869 hCoV-19/England/ALDP-CB410C/2020\|EPI_ISL_761356\|20 | ···················· | ···················· | ···················· | ···················· | ···················· | ···················· |
| AB | 870 hCoV-19/England/MILK-CA662C/2020\|EPI_ISL_761368\|20 | ···················· | ···················· | ···················· | ···················· | ···················· | ···················· |
| AB | 871 hCoV-19/England/MILK-CA6B5A/2020\|EPI_ISL_761380\|20 | ···················· | ···················· | ···················· | ···················· | ···················· | ···················· |
| AB | 872 hCoV-19/England/MILK-CA6EBB/2020\|EPI_ISL_761384\|20 | ···················· | ·A·················· | ···················· | ···················· | ···················· | ··A················· |
| AB | 873 hCoV-19/England/MILK-CAA0DF/2020\|EPI_ISL_761389\|20 | ···················· | ···················· | ···················· | ···················· | ···················· | ···················· |

| | | | | | | | |
|---|---|---|---|---|---|---|---|
| AB | 874 hCoV-19/England/CAMC-C82125/2020\|EPI_ISL_762600\|20 | ···················· | ···················· | ···················· | ···················· | ···················· | ···················· |
| AB | 875 hCoV-19/England/CAMC-C836CB/2020\|EPI_ISL_762603\|20 | ···················· | ···················· | ···················· | ···················· | ···················· | ···················· |
| AB | 876 hCoV-19/England/ALDP-C8A266/2020\|EPI_ISL_762606\|20 | ···················· | ···················· | ···················· | ···················· | ···················· | ···················· |
| AB | 877 hCoV-19/England/CAMC-C83C6F/2020\|EPI_ISL_762611\|20 | ···················· | ···················· | ···················· | ···················· | ···················· | ···················· |
| AB | 878 hCoV-19/England/QEUH-C7CB20/2020\|EPI_ISL_762614\|20 | ···················· | ···················· | ···················· | ···················· | ···················· | ···················· |
| AB | 879 hCoV-19/England/CAMC-C82046/2020\|EPI_ISL_762619\|20 | ···················· | ···················· | ···················· | ···················· | ···················· | ···················· |
| AB | 880 hCoV-19/England/QEUH-C7C6F2/2020\|EPI_ISL_762622\|20 | ···················· | ···················· | ···················· | ···················· | ···················· | ···················· |
| AB | 881 hCoV-19/England/CAMC-C81E2F/2020\|EPI_ISL_762624\|20 | ···················· | ···················· | ···················· | ···················· | ···················· | ···················· |
| AB | 882 hCoV-19/England/CAMC-C807DA/2020\|EPI_ISL_762632\|20 | ···················· | ···················· | ···················· | ···················· | ···················· | ···················· |
| AB | 883 hCoV-19/England/CAMC-C81DE6/2020\|EPI_ISL_762637\|20 | ···················· | ···················· | ···················· | ···················· | ···················· | ···················· |
| AB | 884 hCoV-19/England/CAMC-C80A3B/2020\|EPI_ISL_762677\|20 | ···················· | ···················· | ···················· | ···················· | ···················· | ···················· |
| AB | 885 hCoV-19/England/ALDP-C889DC/2020\|EPI_ISL_762691\|20 | ···················· | ···················· | ···················· | ···················· | ···················· | ···················· |
| AB | 886 hCoV-19/England/ALDP-C8A0E4/2020\|EPI_ISL_762693\|20 | ···················· | ·A·················· | ···················· | ···················· | ···················· | ··A················· |
| AB | 887 hCoV-19/England/CAMC-C80A2C/2020\|EPI_ISL_762699\|20 | ···················· | ···················· | ···················· | ···················· | ···················· | ···················· |
| AB | 888 hCoV-19/England/205141686/2020\|EPI_ISL_766223\|2020 | ···················· | ···················· | ···················· | ···················· | ···················· | ···················· |
| AB | 889 hCoV-19/England/205141823/2020\|EPI_ISL_766224\|2020 | ···················· | ···················· | ···················· | ···················· | ···················· | ···················· |
| AB | 890 hCoV-19/England/205140697/2020\|EPI_ISL_766229\|2020 | ···················· | ···················· | ···················· | ···················· | ···················· | ···················· |
| AB | 891 hCoV-19/England/205090715/2020\|EPI_ISL_766230\|2020 | ···················· | ···················· | ···················· | ···················· | ···················· | ···················· |
| AB | 893 hCoV-19/England/205040389/2020\|EPI_ISL_766246\|2020 | ···················· | ···················· | ···················· | ···················· | ···················· | ···················· |
| AB | 895 hCoV-19/England/205100458/2020\|EPI_ISL_766248\|2020 | ···················· | ···················· | ···················· | ···················· | ···················· | ···················· |
| AB | 896 hCoV-19/England/205100459/2020\|EPI_ISL_766249\|2020 | ···················· | ···················· | ···················· | ···················· | ···················· | ···················· |
| AB | 897 hCoV-19/USA/NY-Wadsworth-289288-01/2020\|EPI_ISL_76 | ···················· | ···················· | ···················· | ···················· | ···················· | ···················· |
| AB | 898 hCoV-19/USA/NY-Wadsworth-289459-01/2020\|EPI_ISL_76 | ···················· | ···················· | ···················· | ···················· | ···················· | ···················· |
| AB | 899 hCoV-19/USA/NY-Wadsworth-289753-01/2020\|EPI_ISL_76 | ···················· | ···················· | ···················· | ···················· | ···················· | ···················· |
| AB | 900 hCoV-19/USA/NY-Wadsworth-289756-01/2020\|EPI_ISL_76 | ···················· | ·A·················· | ···················· | ···················· | ···················· | ···················· |
| AB | 901 hCoV-19/USA/NY-Wadsworth-289757-01/2020\|EPI_ISL_76 | ···················· | ·A·················· | ···················· | ···················· | ···················· | ···················· |
| AB | 902 hCoV-19/USA/NY-Wadsworth-289876-01/2020\|EPI_ISL_76 | ···················· | ···················· | ···················· | ···················· | ···················· | ···················· |
| AB | 903 hCoV-19/USA/NY-Wadsworth-289888-01/2020\|EPI_ISL_76 | ···················· | ·A·················· | ···················· | ···················· | ···················· | ···················· |
| AB | 904 hCoV-19/USA/NY-Wadsworth-289892-01/2020\|EPI_ISL_76 | ···················· | ···················· | ···················· | ···················· | ···················· | ···················· |
| AB | 905 hCoV-19/USA/NY-Wadsworth-290703-01/2020\|EPI_ISL_76 | ···················· | ·············A······ | ···················· | ···················· | ···················· | ···················· |
| AB | 906 hCoV-19/USA/NY-Wadsworth-290705-01/2020\|EPI_ISL_76 | ···················· | ·A·················· | ···················· | ···················· | ···················· | ···················· |
| AB | 907 hCoV-19/USA/NY-Wadsworth-290707-01/2020\|EPI_ISL_76 | ···················· | ·············A······ | ···················· | ···················· | ···················· | ···················· |
| AB | 908 hCoV-19/USA/NY-Wadsworth-290708-01/2020\|EPI_ISL_76 | ···················· | ·············A······ | ···················· | ···················· | ···················· | ···················· |
| AB | 909 hCoV-19/USA/NY-Wadsworth-290709-01/2020\|EPI_ISL_76 | ···················· | ·A·················· | ···················· | ···················· | ···················· | ···················· |
| AB | 910 hCoV-19/USA/NY-Wadsworth-290710-01/2020\|EPI_ISL_76 | ···················· | ·············A······ | ···················· | ···················· | ···················· | ···················· |
| AB | 911 hCoV-19/USA/NY-Wadsworth-290711-01/2020\|EPI_ISL_76 | ···················· | ···················· | ···················· | ···················· | ···················· | ···················· |
| AB | 912 hCoV-19/USA/NY-Wadsworth-290712-01/2020\|EPI_ISL_76 | ···················· | ·A·················· | ···················· | ···················· | ···················· | ···················· |
| AB | 913 hCoV-19/USA/NY-Wadsworth-290714-01/2020\|EPI_ISL_76 | ···················· | ·············A······ | ···················· | ···················· | ···················· | ···················· |
| AB | 914 hCoV-19/USA/NY-Wadsworth-290717-01/2020\|EPI_ISL_76 | ···················· | ···················· | ···················· | ···················· | ···················· | ···················· |
| AB | 915 hCoV-19/USA/NY-Wadsworth-290718-01/2020\|EPI_ISL_76 | ···················· | ···················· | ···················· | ···················· | ···················· | ···················· |
| AB | 916 hCoV-19/USA/NY-Wadsworth-290719-01/2020\|EPI_ISL_76 | ···················· | ·A·················· | ···················· | ···················· | ···················· | ···················· |
| AB | 917 hCoV-19/USA/NY-Wadsworth-290720-01/2020\|EPI_ISL_76 | ···················· | ·············A······ | ···················· | ···················· | ···················· | ···················· |
| AB | 918 hCoV-19/USA/NY-Wadsworth-290723-01/2020\|EPI_ISL_76 | ···················· | ·············A······ | ···················· | ···················· | ···················· | ···················· |
| AB | 919 hCoV-19/USA/NY-Wadsworth-290725-01/2020\|EPI_ISL_76 | ···················· | ·A·················· | ···················· | ···················· | ···················· | ···················· |
| AB | 920 hCoV-19/USA/NY-Wadsworth-290726-01/2020\|EPI_ISL_76 | ···················· | ···················· | ···················· | ···················· | ···················· | ···················· |
| AB | 921 hCoV-19/USA/NY-Wadsworth-290727-01/2020\|EPI_ISL_76 | ···················· | ·A·················· | ···················· | ···················· | ···················· | ···················· |
| AB | 922 hCoV-19/USA/NY-Wadsworth-290728-01/2020\|EPI_ISL_76 | ···················· | ·············A······ | ···················· | ···················· | ···················· | ···················· |
| AB | 923 hCoV-19/USA/NY-Wadsworth-290729-01/2020\|EPI_ISL_76 | ···················· | ·A·················· | ···················· | ···················· | ···················· | ···················· |
| AB | 924 hCoV-19/USA/NY-Wadsworth-290731-01/2020\|EPI_ISL_76 | ···················· | ·A·················· | ···················· | ···················· | ···················· | ···················· |
| AB | 925 hCoV-19/USA/NY-Wadsworth-290732-01/2020\|EPI_ISL_76 | ···················· | ·A·················· | ···················· | ···················· | ···················· | ···················· |
| AB | 926 hCoV-19/USA/NY-Wadsworth-290734-01/2020\|EPI_ISL_76 | ···················· | ···················· | ···················· | ···················· | ···················· | ···················· |
| AB | 927 hCoV-19/USA/NY-Wadsworth-290735-01/2020\|EPI_ISL_76 | ···················· | ·A·················· | ···················· | ···················· | ···················· | ···················· |
| AB | 928 hCoV-19/USA/NY-Wadsworth-290736-01/2020\|EPI_ISL_76 | ···················· | ···················· | ···················· | ···················· | ···················· | ···················· |
| AB | 929 hCoV-19/USA/NY-Wadsworth-290738-01/2020\|EPI_ISL_76 | ···················· | ···················· | ···················· | ···················· | ···················· | ···················· |
| AB | 930 hCoV-19/USA/NY-Wadsworth-290741-01/2020\|EPI_ISL_76 | ···················· | ···················· | ···················· | ···················· | ···················· | ···················· |
| AB | 931 hCoV-19/USA/NY-Wadsworth-290742-01/2020\|EPI_ISL_76 | ···················· | ·A·················· | ···················· | ···················· | ···················· | ···················· |
| AB | 932 hCoV-19/USA/NY-Wadsworth-290744-01/2020\|EPI_ISL_76 | ···················· | ···················· | ···················· | ···················· | ···················· | ···················· |
| AB | 933 hCoV-19/USA/NY-Wadsworth-290745-01/2020\|EPI_ISL_76 | ···················· | ·A·················· | ···················· | ···················· | ···················· | ···················· |
| AB | 934 hCoV-19/USA/NY-Wadsworth-290746-01/2020\|EPI_ISL_76 | ···················· | ·············A······ | ···················· | ···················· | ···················· | ···················· |
| AB | 935 hCoV-19/USA/NY-Wadsworth-290747-01/2020\|EPI_ISL_76 | ···················· | ···················· | ···················· | ···················· | ···················· | ···················· |
| AB | 936 hCoV-19/USA/NY-Wadsworth-290749-01/2020\|EPI_ISL_76 | ···················· | ···················· | ···················· | ···················· | ···················· | ···················· |
| AB | 937 hCoV-19/USA/NY-Wadsworth-290751-01/2020\|EPI_ISL_76 | ···················· | ···················· | ···················· | ···················· | ···················· | ···················· |
| AB | 938 hCoV-19/USA/NY-Wadsworth-290752-01/2020\|EPI_ISL_76 | ···················· | ···················· | ···················· | ···················· | ···················· | ···················· |
| AB | 939 hCoV-19/USA/NY-Wadsworth-290753-01/2020\|EPI_ISL_76 | ···················· | ···················· | ···················· | ···················· | ···················· | ···················· |
| AB | 940 hCoV-19/USA/NY-Wadsworth-290754-01/2020\|EPI_ISL_76 | ···················· | ·A·················· | ···················· | ···················· | ···················· | ···················· |
| AB | 941 hCoV-19/USA/NY-Wadsworth-290755-01/2020\|EPI_ISL_76 | ···················· | ···················· | ···················· | ···················· | ···················· | ···················· |
| AB | 942 hCoV-19/USA/NY-Wadsworth-290757-01/2020\|EPI_ISL_76 | ···················· | ···················· | ···················· | ···················· | ···················· | ···················· |
| AB | 943 hCoV-19/USA/NY-Wadsworth-290758-01/2020\|EPI_ISL_76 | ···················· | ·A·················· | ···················· | ···················· | ···················· | ···················· |
| AB | 944 hCoV-19/USA/NY-Wadsworth-290760-01/2020\|EPI_ISL_76 | ···················· | ·A·················· | ···················· | ···················· | ···················· | ···················· |
| AB | 945 hCoV-19/USA/NY-Wadsworth-229080-01/2020\|EPI_ISL_76 | ···················· | ···················· | ···················· | ···················· | ···················· | ···················· |
| AB | 946 hCoV-19/USA/NY-Wadsworth-290762-01/2020\|EPI_ISL_76 | ···················· | ·A·················· | ···················· | ···················· | ···················· | ···················· |
| AB | 947 hCoV-19/England/205060371/2020\|EPI_ISL_768803\|2020 | ···················· | ···················· | ···················· | ···················· | ···················· | ···················· |
| AB | 948 hCoV-19/England/205090725/2020\|EPI_ISL_768809\|2020 | ···················· | ···················· | ···················· | ···················· | ···················· | ···················· |
| AB | 949 hCoV-19/England/205090727/2020\|EPI_ISL_768811\|2020 | ···················· | ···················· | ···················· | ···················· | ···················· | ···················· |
| AB | 950 hCoV-19/England/205090731/2020\|EPI_ISL_768812\|2020 | ···················· | ···················· | ···················· | ···················· | ···················· | ···················· |
| AB | 951 hCoV-19/England/205090732/2020\|EPI_ISL_768813\|2020 | ···················· | ···················· | ···················· | ···················· | ···················· | ···················· |
| AB | 952 hCoV-19/England/205090734/2020\|EPI_ISL_768814\|2020 | ···················· | ···················· | ···················· | ···················· | ···················· | ···················· |
| AB | 953 hCoV-19/Morocco/FMP-34/2020\|EPI_ISL_768832\|2020-12 | ···················· | ···················· | ···················· | ···················· | ···················· | ···················· |
| AB | 954 hCoV-19/Ireland/D-NVRL-AIIDV1334v1/2020\|EPI_ISL_76 | ···················· | ···················· | ···················· | ···················· | ···················· | ···················· |
| AB | 955 hCoV-19/Morocco/FMP-35/2020\|EPI_ISL_768834\|2020-12 | T··············A···· | ···················· | ···················· | ···················· | ···················· | ···················· |

| | | | | | | | | | |
|---|---|---|---|---|---|---|---|---|---|
| AB | 956 | hCoV-19/Morocco/FMP-36/2020\|EPI_ISL_768835\|2020-12 | .................... | .................... | .................... | .................... | .................... | .................... | |
| AB | 957 | hCoV-19/Morocco/FMP-41/2020\|EPI_ISL_768840\|2020-12 | .................... | .................... | .................... | .................... | .................... | .................... | |
| AB | 958 | hCoV-19/England/ALDP-CB401E/2020\|EPI_ISL_768842\|20 | .................... | .................... | .................... | .................... | .................... | .................... | |
| AB | 959 | hCoV-19/England/MILK-CAA1DC/2020\|EPI_ISL_768871\|20 | .................... | .................... | .................... | .................... | .................... | .................... | |
| AB | 960 | hCoV-19/England/MILK-CAA109/2020\|EPI_ISL_768883\|20 | .................... | .................... | .................... | .................... | .................... | .................... | |
| AB | 961 | hCoV-19/England/MILK-CAA38B/2020\|EPI_ISL_768888\|20 | .................... | .................... | .................... | .................... | .................... | .................... | |
| AB | 962 | hCoV-19/England/MILK-CAA312/2020\|EPI_ISL_768891\|20 | .................... | .................... | .................... | .................... | .................... | .................... | |
| AB | 963 | hCoV-19/England/ALDP-CB5CFB/2020\|EPI_ISL_768899\|20 | .................... | .................... | .................... | .................... | .................... | .................... | |
| AB | 964 | hCoV-19/Germany/HH-hpi-p1450/2020\|EPI_ISL_775910\|2 | .................... | .................... | .................... | .................... | .................... | .................... | |
| AB | 965 | hCoV-19/Germany/HH-hpi-p1451/2020\|EPI_ISL_775911\|2 | .................... | .................... | .................... | .................... | .................... | .................... | |
| AB | 966 | hCoV-19/Germany/HH-hpi-p1459/2020\|EPI_ISL_775912\|2 | .................... | .................... | .................... | .................... | .................... | .................... | |
| AB | 967 | hCoV-19/Germany/HH-hpi-p1473/2020\|EPI_ISL_775913\|2 | .................... | .................... | .................... | .................... | .................... | .................... | |
| AB | 968 | hCoV-19/Germany/HH-hpi-p1495/2020\|EPI_ISL_775914\|2 | .................... | .................... | .................... | .................... | .................... | .................... | |
| AB | 969 | hCoV-19/Germany/HH-hpi-p1505/2020\|EPI_ISL_775915\|2 | .................... | N---................ | .................... | .................... | .................... | .................... | 1 |
| AB | 970 | hCoV-19/Germany/HH-hpi-p1792/2020\|EPI_ISL_775916\|2 | .................... | .................... | .................... | .................... | .................... | .................... | |
| AB | 971 | hCoV-19/Germany/HH-hpi-p1816/2020\|EPI_ISL_775917\|2 | .................... | .................... | .................... | .................... | .................... | .................... | |
| AB | 972 | hCoV-19/Germany/HH-hpi-p1853/2020\|EPI_ISL_775918\|2 | .................... | .................... | .................... | .................... | .................... | .................... | |
| AB | 973 | hCoV-19/Germany/HH-hpi-p1862/2020\|EPI_ISL_775919\|2 | .................... | .................... | .................... | .................... | .................... | .................... | |
| AB | 974 | hCoV-19/Germany/HH-hpi-p1903/2020\|EPI_ISL_775920\|2 | .................... | .................... | .................... | .................... | .................... | .................... | |
| AB | 976 | hCoV-19/Germany/HH-hpi-p1546/2020\|EPI_ISL_775927\|2 | .................... | .................... | .................... | .................... | .................... | .................... | |
| AB | 977 | hCoV-19/Germany/HH-hpi-p1612/2020\|EPI_ISL_775928\|2 | .................... | .................... | .................... | .................... | .................... | .................... | |
| AB | 978 | hCoV-19/Germany/HH-hpi-p1633/2020\|EPI_ISL_775929\|2 | .................... | .................... | .................... | .................... | .................... | .................... | |
| AB | 979 | hCoV-19/Germany/HH-hpi-p1655/2020\|EPI_ISL_775930\|2 | .................... | .................... | .................... | .................... | .................... | .................... | |
| AB | 980 | hCoV-19/Germany/HH-hpi-p1809/2020\|EPI_ISL_775931\|2 | .................... | .................... | .................... | .................... | .................... | .................... | |
| AB | 981 | hCoV-19/Germany/HH-hpi-p1859/2020\|EPI_ISL_775932\|2 | .................... | .................... | .................... | .................... | .................... | .................... | |
| AB | 982 | hCoV-19/Germany/HH-hpi-p1911/2020\|EPI_ISL_775933\|2 | .................... | .................... | .................... | .................... | .................... | .................... | |
| AB | 983 | hCoV-19/Germany/HH-hpi-p2034/2020\|EPI_ISL_775934\|2 | .................... | .................... | .................... | .................... | .................... | .................... | |
| AB | 984 | hCoV-19/Germany/HH-hpi-p2066/2020\|EPI_ISL_775935\|2 | .................... | .................... | .................... | .................... | .................... | .................... | |
| AB | 986 | hCoV-19/Germany/HH-hpi-p2328/2020\|EPI_ISL_775937\|2 | .................... | .................... | .................... | .................... | .................... | .................... | |
| AB | 987 | hCoV-19/Germany/HH-hpi-p2331/2020\|EPI_ISL_775938\|2 | .................... | .................... | .................... | .................... | .................... | .................... | |
| AB | 989 | hCoV-19/Germany/SH-hpi-p1415/2020\|EPI_ISL_775940\|2 | .................... | .................... | .................... | .................... | .................... | .................... | |
| AB | 990 | hCoV-19/Germany/HH-hpi-p1412/2020\|EPI_ISL_775941\|2 | .................... | .................... | .................... | .................... | .................... | .................... | |
| AB | 991 | hCoV-19/Germany/HH-hpi-p1414/2020\|EPI_ISL_775942\|2 | .................... | .................... | .................... | .................... | .................... | .................... | |
| AB | 992 | hCoV-19/Germany/HH-hpi-p1416/2020\|EPI_ISL_775943\|2 | .................... | .................... | .................... | .................... | .................... | .................... | |
| AB | 993 | hCoV-19/Germany/HH-hpi-p1418/2020\|EPI_ISL_775944\|2 | .................... | .................... | .................... | .................... | .................... | .................... | |
| AB | 994 | hCoV-19/Germany/HH-hpi-p1419/2020\|EPI_ISL_775945\|2 | .................... | .................... | .................... | .................... | .................... | .................... | |
| AB | 995 | hCoV-19/Germany/HH-hpi-p1422/2020\|EPI_ISL_775946\|2 | .................... | .................... | .................... | .................... | .................... | .................... | |
| AB | 996 | hCoV-19/Germany/HH-hpi-p1423/2020\|EPI_ISL_775947\|2 | .................... | .................... | .................... | .................... | .................... | .................... | |
| AB | 997 | hCoV-19/Germany/HH-hpi-p1427/2020\|EPI_ISL_775948\|2 | .................... | .................... | .................... | .................... | .................... | .................... | |
| AB | 998 | hCoV-19/Germany/HH-hpi-p1428/2020\|EPI_ISL_775949\|2 | .................... | .................... | .................... | .................... | .................... | .................... | |
| AB | 999 | hCoV-19/Germany/HH-hpi-p1429/2020\|EPI_ISL_775950\|2 | .................... | .................... | .................... | .................... | .................... | .................... | |
| AB | 1000 | hCoV-19/Germany/HH-hpi-p1430/2020\|EPI_ISL_775951\|2 | .................... | .................... | .................... | .................... | .................... | .................... | |
| AB | 1001 | hCoV-19/Germany/HH-hpi-p1431/2020\|EPI_ISL_775952\|2 | .................... | .................... | .................... | .................... | .................... | .................... | |
| AB | 1002 | hCoV-19/Germany/HH-hpi-p1465/2020\|EPI_ISL_775978\|2 | .................... | .................... | .................... | .................... | .................... | .................... | |
| AB | 1003 | hCoV-19/Germany/HH-hpi-p1468/2020\|EPI_ISL_775980\|2 | .................... | .................... | .................... | .................... | .................... | .................... | |
| AB | 1004 | hCoV-19/Germany/HH-hpi-p1469/2020\|EPI_ISL_775981\|2 | .................... | .................... | .................... | .................... | .................... | .................... | |
| AB | 1005 | hCoV-19/Germany/HH-hpi-p1470/2020\|EPI_ISL_775982\|2 | .................... | .................... | .................... | .................... | .................... | .................... | |
| AB | 1006 | hCoV-19/Germany/HH-hpi-p1471/2020\|EPI_ISL_775983\|2 | .................... | .................... | .................... | .................... | .................... | .................... | |
| AB | 1007 | hCoV-19/Germany/HH-hpi-p1472/2020\|EPI_ISL_775984\|2 | .................... | .................... | .................... | .................... | .................... | .................... | |
| AB | 1008 | hCoV-19/Germany/HH-hpi-p1474/2020\|EPI_ISL_775985\|2 | .................... | .................... | .................... | .................... | .................... | .................... | |
| AB | 1009 | hCoV-19/Germany/HH-hpi-p1487/2020\|EPI_ISL_775997\|2 | .................... | .................... | .................... | .................... | .................... | .................... | |
| AB | 1010 | hCoV-19/Germany/HH-hpi-p1489/2020\|EPI_ISL_775998\|2 | .................... | .................... | .................... | .................... | .................... | .................... | |
| AB | 1011 | hCoV-19/Germany/HH-hpi-p1491/2020\|EPI_ISL_775999\|2 | .................... | .................... | .................... | .................... | .................... | .................... | |
| AB | 1012 | hCoV-19/England/ALDP-CBF480/2020\|EPI_ISL_778190\|20 | .................... | .................... | .................... | .................... | .................... | .................... | |
| AB | 1013 | hCoV-19/England/QEUH-CC3058/2020\|EPI_ISL_778192\|20 | .................... | .................... | .................... | .................... | .................... | .................... | |
| AB | 1014 | hCoV-19/England/QEUH-CC302B/2020\|EPI_ISL_778203\|20 | .................... | .................... | .................... | .................... | .................... | .................... | |
| AB | 1015 | hCoV-19/England/CAMC-CB8A1C/2020\|EPI_ISL_778206\|20 | .................... | .................... | .................... | .................... | .................... | .................... | |
| AB | 1016 | hCoV-19/England/ALDP-CC096F/2020\|EPI_ISL_778207\|20 | .................... | .................... | .................... | .................... | .................... | .................... | |
| AB | 1017 | hCoV-19/England/CAMC-CB751C/2020\|EPI_ISL_778208\|20 | .................... | .................... | .................... | .................... | .................... | .................... | |
| AB | 1018 | hCoV-19/England/CAMC-CB7479/2020\|EPI_ISL_778211\|20 | .................... | .................... | .................... | .................... | .................... | .................... | |
| AB | 1019 | hCoV-19/Wales/ALDP-CC0783/2020\|EPI_ISL_778212\|2020 | .................... | .................... | .................... | .................... | .................... | .................... | |
| AB | 1020 | hCoV-19/England/QEUH-CC2745/2020\|EPI_ISL_778215\|20 | .................... | .................... | .................... | .................... | ....T............... | .................... | |
| AB | 1021 | hCoV-19/England/QEUH-CC2596/2020\|EPI_ISL_778216\|20 | .................... | .................... | .................... | .................... | .................... | .................... | |
| AB | 1022 | hCoV-19/England/QEUH-CC2DAD/2020\|EPI_ISL_778219\|20 | .................... | .................... | .................... | .................... | .................... | .................... | |
| AB | 1023 | hCoV-19/England/CAMC-CB7312/2020\|EPI_ISL_778221\|20 | .................... | .A.................. | .................... | .................... | .................... | ..A................. | |
| AB | 1024 | hCoV-19/England/ALDP-CC07CF/2020\|EPI_ISL_778225\|20 | .................... | .................... | .................... | .................... | .................... | .................... | |
| AB | 1025 | hCoV-19/South Korea/KDCA0314/2020\|EPI_ISL_760100\|2 | .................... | .................... | .................... | .................... | .................... | .................... | |
| AB | 1026 | hCoV-19/South Korea/KDCA0315/2020\|EPI_ISL_760101\|2 | .................... | .................... | .................... | .................... | .................... | .................... | |
| AB | 1027 | hCoV-19/South Korea/KDCA0316/2020\|EPI_ISL_760102\|2 | .................... | .................... | .................... | .................... | .................... | .................... | |
| AB | 1028 | hCoV-19/South Korea/KDCA0317/2020\|EPI_ISL_760103\|2 | .................... | .................... | .................... | .................... | .................... | .................... | |
| AB | 1029 | hCoV-19/South Korea/KDCA0318/2020\|EPI_ISL_760104\|2 | .................... | .................... | .................... | .................... | .................... | .................... | |
| AB | 1030 | hCoV-19/South Korea/KDCA0319/2020\|EPI_ISL_760105\|2 | .................... | .................... | .................... | .................... | .................... | .................... | |
| AB | 1031 | hCoV-19/South Korea/KDCA0320/2020\|EPI_ISL_760106\|2 | .................... | .................... | .................... | .................... | .................... | .................... | |
| AB | 1032 | hCoV-19/South Korea/KDCA0321/2020\|EPI_ISL_760107\|2 | .................... | .................... | .................... | .................... | .................... | .................... | |
| AB | 1033 | hCoV-19/South Korea/KDCA0322/2020\|EPI_ISL_760108\|2 | .................... | .................... | .................... | .................... | .................... | .................... | |
| AB | 1034 | hCoV-19/South Korea/KDCA0324/2020\|EPI_ISL_760110\|2 | .................... | .................... | .................... | .................... | .................... | .................... | |
| AB | 1035 | hCoV-19/South Korea/KDCA0325/2020\|EPI_ISL_760111\|2 | .................... | .................... | .................... | .................... | .................... | .................... | |
| AB | 1036 | hCoV-19/South Korea/KDCA0326/2020\|EPI_ISL_760112\|2 | .................... | .................... | .................... | .................... | .................... | .................... | |
| AB | 1037 | hCoV-19/South Korea/KDCA0327/2020\|EPI_ISL_760113\|2 | .................... | .................... | .................... | .................... | .................... | .................... | |
| AB | 1038 | hCoV-19/South Korea/KDCA0328/2020\|EPI_ISL_760114\|2 | .................... | .................... | .................... | .................... | .................... | .................... | |

| | | | | | | | |
|---|---|---|---|---|---|---|---|
| AB | 1039 hCoV-19/South Korea/KDCA0329/2020\|EPI_ISL_760115\|2 | .................... | .................... | ..................... | .................... | .................... | .................... |
| AB | 1040 hCoV-19/South Korea/KDCA0330/2020\|EPI_ISL_760116\|2 | .................... | .................... | ..................... | .................... | .................... | .................... |
| AB | 1041 hCoV-19/South Korea/KDCA0331/2020\|EPI_ISL_760117\|2 | .................... | .................... | ..................... | .................... | .................... | .................... |
| AB | 1042 hCoV-19/South Korea/KDCA0332/2020\|EPI_ISL_760118\|2 | .................... | .................... | ..................... | .................... | .................... | .................... |
| AB | 1043 hCoV-19/South Korea/KDCA0333/2020\|EPI_ISL_760119\|2 | .................... | .................... | ..................... | .................... | .................... | .................... |
| AB | 1044 hCoV-19/South Korea/KDCA0334/2020\|EPI_ISL_760120\|2 | .................... | .................... | ..................... | .................... | .................... | .................... |
| AB | 1045 hCoV-19/South Korea/KDCA0335/2020\|EPI_ISL_760121\|2 | .................... | .................... | ..................... | .................... | .................... | .................... |
| AB | 1046 hCoV-19/South Korea/KDCA0336/2020\|EPI_ISL_760122\|2 | .................... | .................... | ..................... | .................... | .................... | .................... |
| AB | 1047 hCoV-19/South Korea/KDCA0337/2020\|EPI_ISL_760123\|2 | .................... | .................... | ..................... | .................... | .................... | .................... |
| AB | 1048 hCoV-19/South Korea/KDCA0338/2020\|EPI_ISL_760124\|2 | .................... | .................... | ..................... | .................... | .................... | .................... |
| AB | 1049 hCoV-19/South Korea/KDCA0339/2020\|EPI_ISL_760125\|2 | .................... | .................... | ..................... | .................... | .................... | .................... |
| AB | 1050 hCoV-19/South Korea/KDCA0340/2020\|EPI_ISL_760126\|2 | .................... | .................... | ..................... | .................... | .................... | .................... |
| AB | 1051 hCoV-19/South Korea/KDCA0341/2020\|EPI_ISL_760127\|2 | .................... | .................... | ..................... | .................... | .................... | .................... |
| AB | 1052 hCoV-19/South Korea/KDCA0342/2020\|EPI_ISL_760128\|2 | .................... | .................... | ..................... | .................... | .................... | .................... |
| AB | 1053 hCoV-19/South Korea/KDCA0343/2020\|EPI_ISL_760129\|2 | .................... | .................... | ..................... | .................... | .................... | .................... |
| AB | 1054 hCoV-19/South Korea/KDCA0344/2020\|EPI_ISL_760130\|2 | .................... | .................... | ..................... | .................... | .................... | .................... |
| AB | 1055 hCoV-19/South Korea/KDCA0345/2020\|EPI_ISL_760131\|2 | .................... | .................... | ..................... | .................... | .................... | .................... |
| AB | 1056 hCoV-19/South Korea/KDCA0346/2020\|EPI_ISL_760132\|2 | .................... | .................... | ..................... | .................... | .................... | .................... |
| AB | 1057 hCoV-19/South Korea/KDCA0348/2020\|EPI_ISL_760133\|2 | .................... | .................... | ..................... | .................... | .................... | .................... |
| AB | 1058 hCoV-19/South Korea/KDCA0349/2020\|EPI_ISL_760134\|2 | .................... | .................... | ..................... | .................... | .................... | .................... |
| AB | 1059 hCoV-19/South Korea/KDCA0350/2020\|EPI_ISL_760135\|2 | .................... | .................... | ..................... | .................... | .................... | .................... |
| AB | 1060 hCoV-19/South Korea/KDCA0351/2020\|EPI_ISL_760136\|2 | .................... | .................... | ..................... | .................... | .................... | .................... |
| AB | 1061 hCoV-19/South Korea/KDCA0352/2020\|EPI_ISL_760137\|2 | .................... | .................... | ..................... | .................... | .................... | .................... |
| AB | 1062 hCoV-19/South Korea/KDCA0353/2020\|EPI_ISL_760138\|2 | .................... | .................... | ..................... | .................... | .................... | .................... |
| AB | 1063 hCoV-19/South Korea/KDCA0354/2020\|EPI_ISL_760139\|2 | .................... | .................... | ..................... | .................... | .................... | .................... |
| AB | 1064 hCoV-19/South Korea/KDCA0355/2020\|EPI_ISL_760140\|2 | .................... | .................... | ..................... | .................... | .................... | .................... |
| AB | 1065 hCoV-19/South Korea/KDCA0356/2020\|EPI_ISL_760141\|2 | .................... | .................... | ..................... | .................... | .................... | .................... |
| AB | 1066 hCoV-19/South Korea/KDCA0357/2020\|EPI_ISL_760142\|2 | .................... | .................... | ..................... | .................... | .................... | .................... |
| AB | 1067 hCoV-19/South Korea/KDCA0358/2020\|EPI_ISL_760143\|2 | .................... | .................... | ..................... | .................... | .................... | .................... |
| AB | 1068 hCoV-19/South Korea/KDCA0359/2020\|EPI_ISL_760144\|2 | .................... | .................... | ..................... | .................... | .................... | .................... |
| AB | 1069 hCoV-19/South Korea/KDCA0360/2020\|EPI_ISL_760145\|2 | .................... | .................... | ..................... | .................... | .................... | .................... |
| AB | 1070 hCoV-19/South Korea/KDCA0361/2020\|EPI_ISL_760146\|2 | .................... | .................... | ..................... | .................... | .................... | .................... |
| AB | 1071 hCoV-19/South Korea/KDCA0363/2020\|EPI_ISL_760147\|2 | .................... | .................... | ..................... | .................... | .................... | .................... |
| AB | 1072 hCoV-19/South Korea/KDCA0364/2020\|EPI_ISL_760148\|2 | .................... | .................... | ..................... | .................... | .................... | .................... |
| AB | 1073 hCoV-19/South Korea/KDCA0365/2020\|EPI_ISL_760149\|2 | .................... | .................... | ..................... | .................... | .................... | .................... |
| AB | 1074 hCoV-19/South Korea/KDCA0366/2020\|EPI_ISL_760150\|2 | .................... | .................... | ..................... | .................... | .................... | .................... |
| AB | 1075 hCoV-19/South Korea/KDCA0367/2020\|EPI_ISL_760151\|2 | .................... | .................... | ..................... | .................... | .................... | .................... |
| AB | 1076 hCoV-19/South Korea/KDCA0368/2020\|EPI_ISL_760152\|2 | .................... | .................... | ..................... | .................... | .................... | .................... |
| AB | 1077 hCoV-19/South Korea/KDCA0369/2020\|EPI_ISL_760153\|2 | .................... | .................... | ..................... | .................... | .................... | .................... |
| AB | 1078 hCoV-19/South Korea/KDCA0370/2020\|EPI_ISL_760154\|2 | .................... | .................... | ..................... | .................... | .................... | .................... |
| AB | 1079 hCoV-19/South Korea/KDCA0371/2020\|EPI_ISL_760155\|2 | .................... | .................... | ..................... | .................... | .................... | .................... |
| AB | 1080 hCoV-19/South Korea/KDCA0372/2020\|EPI_ISL_760156\|2 | .................... | .................... | ..................... | .................... | .................... | .................... |
| AB | 1081 hCoV-19/South Korea/KDCA0380/2020\|EPI_ISL_760164\|2 | .................... | .................... | ..................... | .................... | .................... | .................... |
| AB | 1082 hCoV-19/South Korea/KDCA0381/2020\|EPI_ISL_760165\|2 | .................... | .................... | ..................... | .................... | .................... | .................... |
| AB | 1083 hCoV-19/South Korea/KDCA0382/2020\|EPI_ISL_760166\|2 | .................... | .................... | ..................... | .................... | .................... | .................... |
| AB | 1084 hCoV-19/South Korea/KDCA0383/2020\|EPI_ISL_760167\|2 | .................... | .................... | ..................... | .................... | .................... | .................... |
| AB | 1085 hCoV-19/South Korea/KDCA0406/2020\|EPI_ISL_760190\|2 | .................... | .................... | ..................... | .................... | .................... | .................... |
| AB | 1086 hCoV-19/South Korea/KDCA0407/2020\|EPI_ISL_760191\|2 | .................... | .................... | ..................... | .................... | .................... | .................... |
| AB | 1087 hCoV-19/South Korea/KDCA0408/2020\|EPI_ISL_760192\|2 | .................... | .................... | ..................... | .................... | .................... | .................... |
| AB | 1088 hCoV-19/South Korea/KDCA0409/2020\|EPI_ISL_760193\|2 | .................... | .................... | ..................... | .................... | .................... | .................... |
| AB | 1089 hCoV-19/South Korea/KDCA0410/2020\|EPI_ISL_760194\|2 | .................... | .................... | ..................... | .................... | .................... | .................... |
| AB | 1090 hCoV-19/South Korea/KDCA0411/2020\|EPI_ISL_760195\|2 | .................... | .................... | ..................... | .................... | .................... | .................... |
| AB | 1091 hCoV-19/South Korea/KDCA0412/2020\|EPI_ISL_760196\|2 | .................... | .................... | ..................... | .................... | .................... | .................... |
| AB | 1092 hCoV-19/South Korea/KDCA0413/2020\|EPI_ISL_760197\|2 | .................... | .................... | ..................... | .................... | .................... | .................... |
| AB | 1093 hCoV-19/South Korea/KDCA0414/2020\|EPI_ISL_760198\|2 | .................... | .................... | ..................... | .................... | .................... | .................... |
| AB | 1094 hCoV-19/South Korea/KDCA0415/2020\|EPI_ISL_760199\|2 | .................... | .................... | ..................... | .................... | .................... | .................... |
| AB | 1095 hCoV-19/England/MILK-CA6853/2020\|EPI_ISL_761406\|20 | .................... | .................... | ..................... | .................... | .................... | .................... |
| AB | 1096 hCoV-19/England/MILK-CA6C66/2020\|EPI_ISL_761408\|20 | .................... | .................... | ..................... | .................... | .................... | .................... |
| AB | 1097 hCoV-19/England/MILK-CA6A02/2020\|EPI_ISL_761410\|20 | .................... | .................... | ..................... | .................... | .................... | .................... |
| AB | 1098 hCoV-19/England/MILK-CAA02A/2020\|EPI_ISL_761411\|20 | .................... | .................... | ..................... | .................... | ......T............. | .................... |
| AB | 1099 hCoV-19/England/MILK-CA71EE/2020\|EPI_ISL_761413\|20 | .................... | .................... | ..................... | .................... | .................... | .................... |
| AB | 1100 hCoV-19/England/MILK-CA6FF4/2020\|EPI_ISL_761416\|20 | .................... | .................... | ..................... | .................... | ......T............. | .................... |
| AB | 1101 hCoV-19/England/MILK-CA6DAF/2020\|EPI_ISL_761417\|20 | .................... | .................... | ..................... | .................... | .................... | .................... |
| AB | 1102 hCoV-19/England/MILK-CA5D64/2020\|EPI_ISL_761429\|20 | .................... | .................... | ..................... | .................... | .................... | .................... |
| AB | 1103 hCoV-19/England/MILK-CA605B/2020\|EPI_ISL_761432\|20 | .................... | .................... | ..................... | .................... | .................... | .................... |
| AB | 1104 hCoV-19/England/MILK-CA5C85/2020\|EPI_ISL_761435\|20 | .................... | .................... | ..................... | .................... | ......T............. | .................... |
| AB | 1105 hCoV-19/England/MILK-CA6941/2020\|EPI_ISL_761439\|20 | .................... | .................... | ..................... | .................... | .................... | .................... |
| AB | 1106 hCoV-19/England/MILK-CA652F/2020\|EPI_ISL_761441\|20 | .................... | .................... | ..................... | .................... | .................... | .................... |
| AB | 1107 hCoV-19/England/MILK-CA68BD/2020\|EPI_ISL_761446\|20 | .................... | .................... | ..................... | .................... | .................... | .................... |
| AB | 1108 hCoV-19/England/MILK-CA6880/2020\|EPI_ISL_761447\|20 | .................... | .................... | ..................... | .................... | .................... | .................... |
| AB | 1109 hCoV-19/England/MILK-CA9F97/2020\|EPI_ISL_761452\|20 | .................... | .................... | ..................... | .................... | .................... | .................... |
| AB | 1110 hCoV-19/England/MILK-CAA0B1/2020\|EPI_ISL_761454\|20 | .................... | .................... | ..................... | .................... | .................... | .................... |
| AB | 1111 hCoV-19/England/MILK-CA639E/2020\|EPI_ISL_761464\|20 | .................... | .................... | ..................... | .................... | .................... | .................... |
| AB | 1112 hCoV-19/England/MILK-CA62CE/2020\|EPI_ISL_761475\|20 | .................... | .................... | ..................... | .................... | .................... | .................... |
| AB | 1113 hCoV-19/England/MILK-CA6D63/2020\|EPI_ISL_761478\|20 | .................... | .................... | ..................... | .................... | .................... | .................... |
| AB | 1114 hCoV-19/England/MILK-CA70B4/2020\|EPI_ISL_761482\|20 | .................... | .................... | ..................... | .................... | .................... | .................... |
| AB | 1115 hCoV-19/England/MILK-CA6862/2020\|EPI_ISL_761495\|20 | .................... | .................... | ..................... | .................... | .................... | .................... |
| AB | 1116 hCoV-19/England/MILK-CA70F0/2020\|EPI_ISL_761497\|20 | .................... | .................... | ..................... | .................... | .................... | .................... |
| AB | 1117 hCoV-19/England/ALDP-C8874B/2020\|EPI_ISL_762706\|20 | .................... | .................... | ..................... | .................... | .................... | .................... |
| AB | 1118 hCoV-19/England/CAMC-C82161/2020\|EPI_ISL_762716\|20 | .................... | .................... | ..................... | .................... | .................... | .................... |

| | | | | | | | | |
|---|---|---|---|---|---|---|---|---|
| AB | 1119 | hCoV-19/England/CAMC-C80AA4/2020\|EPI_ISL_762717\|20 | ···················· | ···················· | ···················· | ···················· | ···················· | ···················· |
| AB | 1120 | hCoV-19/England/CAMC-C83D20/2020\|EPI_ISL_762723\|20 | ···················· | ···················· | ···················· | ···················· | ···················· | ···················· |
| AB | 1121 | hCoV-19/England/CAMC-C836E9/2020\|EPI_ISL_762725\|20 | ···················· | ···················· | ···················· | ···················· | ···················· | ···················· |
| AB | 1122 | hCoV-19/England/CAMC-C8079E/2020\|EPI_ISL_762727\|20 | ···················· | ···················· | ···················· | ···················· | ···················· | ···················· |
| AB | 1123 | hCoV-19/England/CAMC-C82143/2020\|EPI_ISL_762729\|20 | ···················· | ···················· | ···················· | ···················· | ···················· | ···················· |
| AB | 1124 | hCoV-19/England/ALDP-C8B56C/2020\|EPI_ISL_762733\|20 | ···················· | ···················· | ···················· | ···················· | ···················· | ···················· |
| AB | 1125 | hCoV-19/England/CAMC-C8200A/2020\|EPI_ISL_762737\|20 | ···················· | ···················· | ···················· | ···················· | ···················· | ···················· |
| AB | 1126 | hCoV-19/England/CAMC-C8369E/2020\|EPI_ISL_762738\|20 | ···················· | ···················· | ···················· | ···················· | ······T············· | ···················· |
| AB | 1127 | hCoV-19/England/QEUH-C7CA8D/2020\|EPI_ISL_762739\|20 | ···················· | ···················· | ···················· | ···················· | ···················· | ···················· |
| AB | 1128 | hCoV-19/England/CAMC-C82ED3/2020\|EPI_ISL_762745\|20 | ···················· | ···················· | ···················· | ···················· | ···················· | ···················· |
| AB | 1129 | hCoV-19/England/QEUH-C7C698/2020\|EPI_ISL_762746\|20 | ···················· | ···················· | ···················· | ···················· | ···················· | ···················· |
| AB | 1130 | hCoV-19/England/CAMC-C80AFF/2020\|EPI_ISL_762747\|20 | ···················· | ···················· | ···················· | ···················· | ···················· | ···················· |
| AB | 1131 | hCoV-19/England/CAMC-C836F8/2020\|EPI_ISL_762751\|20 | ···················· | ···················· | ···················· | ···················· | ···················· | ···················· |
| AB | 1132 | hCoV-19/England/QEUH-C7C6B6/2020\|EPI_ISL_762760\|20 | ···················· | ·A·················· | ···················· | ···················· | ···················· | ···A················ |
| AB | 1133 | hCoV-19/England/CAMC-C805B2/2020\|EPI_ISL_762762\|20 | ···················· | ···················· | ···················· | ···················· | ···················· | ···················· |
| AB | 1134 | hCoV-19/England/ALDP-C8873C/2020\|EPI_ISL_762763\|20 | ···················· | ···················· | ···················· | ···················· | ···················· | ···················· |
| AB | 1135 | hCoV-19/England/QEUH-C7D9CB/2020\|EPI_ISL_762773\|20 | ···················· | ···················· | ···················· | ···················· | ···················· | ···················· |
| AB | 1136 | hCoV-19/England/CAMC-C80B1A/2020\|EPI_ISL_762784\|20 | ···················· | ···················· | ···················· | ···················· | ···················· | ···················· |
| AB | 1137 | hCoV-19/England/ALDP-C889BE/2020\|EPI_ISL_762789\|20 | ···················· | ·A·················· | ···················· | ···················· | ···················· | ···A················ |
| AB | 1138 | hCoV-19/England/CAMC-C82477/2020\|EPI_ISL_762792\|20 | ···················· | ···················· | ···················· | ···················· | ······T············· | ···················· |
| AB | 1139 | hCoV-19/England/CAMC-C838F2/2020\|EPI_ISL_762795\|20 | ···················· | ···················· | ···················· | ···················· | ···················· | ···················· |
| AB | 1140 | hCoV-19/England/CAMC-C8390E/2020\|EPI_ISL_762797\|20 | ···················· | ···················· | ···················· | ···················· | ···················· | ···················· |
| AB | 1142 | hCoV-19/England/205181889/2020\|EPI_ISL_766251\|2020 | ···················· | ···················· | ···················· | ···················· | ···················· | ···················· |
| AB | 1143 | hCoV-19/England/205181892/2020\|EPI_ISL_766252\|2020 | ···················· | ···················· | ···················· | ···················· | ···················· | ···················· |
| AB | 1144 | hCoV-19/England/205181910/2020\|EPI_ISL_766253\|2020 | ···················· | ···················· | ···················· | ···················· | ···················· | ···················· |
| AB | 1145 | hCoV-19/England/205181914/2020\|EPI_ISL_766254\|2020 | ···················· | ···················· | ···················· | ···················· | ···················· | ···················· |
| AB | 1146 | hCoV-19/England/205260322/2020\|EPI_ISL_766255\|2020 | ···················· | ···················· | ···················· | ···················· | ···················· | ···················· |
| AB | 1147 | hCoV-19/England/205260364/2020\|EPI_ISL_766256\|2020 | ···················· | ···················· | ···················· | ···················· | ···················· | ···················· |
| AB | 1148 | hCoV-19/England/205260368/2020\|EPI_ISL_766257\|2020 | ···················· | ···················· | ···················· | ···················· | ···················· | ···················· |
| AB | 1149 | hCoV-19/England/205260372/2020\|EPI_ISL_766258\|2020 | ···················· | ···················· | ···················· | ···················· | ···················· | ···················· |
| AB | 1150 | hCoV-19/England/201500413/2020\|EPI_ISL_766259\|2020 | ···················· | ···················· | ···················· | ···················· | ···················· | ···················· |
| AB | 1151 | hCoV-19/England/205100235/2020\|EPI_ISL_766260\|2020 | ···················· | ·A·················· | ···················· | ···················· | ···················· | ···A················ |
| AB | 1152 | hCoV-19/England/204620365/2020\|EPI_ISL_766263\|2020 | ···················· | ···················· | ···················· | ···················· | ···················· | ···················· |
| AB | 1153 | hCoV-19/England/204621636/2020\|EPI_ISL_766264\|2020 | ···················· | ···················· | ···················· | ···················· | ···················· | ···················· |
| AB | 1154 | hCoV-19/England/204661390/2020\|EPI_ISL_766265\|2020 | ···················· | ···················· | ···················· | ···················· | ······T············· | ···················· |
| AB | 1155 | hCoV-19/England/204661395/2020\|EPI_ISL_766266\|2020 | ···················· | ···················· | ···················· | ···················· | ······T············· | ···················· |
| AB | 1156 | hCoV-19/England/204680331/2020\|EPI_ISL_766267\|2020 | ···················· | ·A·················· | ···················· | ···················· | ···················· | ···A················ |
| AB | 1157 | hCoV-19/England/204690821/2020\|EPI_ISL_766268\|2020 | ···················· | ···················· | ···················· | ···················· | ···················· | ···················· |
| AB | 1159 | hCoV-19/England/204691686/2020\|EPI_ISL_766270\|2020 | ···················· | ···················· | ···················· | ···················· | ···················· | ···················· |
| AB | 1161 | hCoV-19/England/204720852/2020\|EPI_ISL_766272\|2020 | ···················· | ·A·······A·········· | ···················· | ···················· | ···················· | ···················· |
| AB | 1165 | hCoV-19/England/204781284/2020\|EPI_ISL_766276\|2020 | ···················· | ···················· | ···················· | ···················· | ······T············· | ···················· |
| AB | 1166 | hCoV-19/England/204840067/2020\|EPI_ISL_766277\|2020 | ···················· | ···················· | ···················· | ···················· | ···················· | ···················· |
| AB | 1168 | hCoV-19/England/204900615/2020\|EPI_ISL_766279\|2020 | ···················· | ···················· | ···················· | ···················· | ···················· | ···················· |
| AB | 1169 | hCoV-19/England/205000383/2020\|EPI_ISL_766280\|2020 | ···················· | ···················· | ···················· | ···················· | ···················· | ···················· |
| AB | 1170 | hCoV-19/England/205020757/2020\|EPI_ISL_766281\|2020 | ···················· | ···················· | ···················· | ···················· | ···················· | ···················· |
| AB | 1171 | hCoV-19/England/205022338/2020\|EPI_ISL_766282\|2020 | ···················· | ···················· | ···················· | ···················· | ···················· | ···················· |
| AB | 1172 | hCoV-19/England/205041299/2020\|EPI_ISL_766283\|2020 | ···················· | ···················· | ···················· | ···················· | ···················· | ···················· |
| AB | 1173 | hCoV-19/England/205041306/2020\|EPI_ISL_766284\|2020 | ···················· | ···················· | ···················· | ···················· | ···················· | ···················· |
| AB | 1174 | hCoV-19/England/205041398/2020\|EPI_ISL_766285\|2020 | ···················· | ·········T·········· | ···················· | ···················· | ···················· | ···················· |
| AB | 1175 | hCoV-19/England/205041400/2020\|EPI_ISL_766286\|2020 | ···················· | ···················· | ···················· | ···················· | ···················· | ···················· |
| AB | 1176 | hCoV-19/England/205041401/2020\|EPI_ISL_766287\|2020 | ···················· | ·A·················· | ···················· | ···················· | ···················· | ···A················ |
| AB | 1177 | hCoV-19/England/205060952/2020\|EPI_ISL_766288\|2020 | ···················· | ···················· | ···················· | ···················· | ···················· | ···················· |
| AB | 1178 | hCoV-19/England/205060997/2020\|EPI_ISL_766289\|2020 | ···················· | ···················· | ···················· | ···················· | ···················· | ···················· |
| AB | 1179 | hCoV-19/England/205081143/2020\|EPI_ISL_766290\|2020 | ···················· | ···················· | ···················· | ···················· | ···················· | ···················· |
| AB | 1180 | hCoV-19/England/205081167/2020\|EPI_ISL_766291\|2020 | ···················· | ···················· | ···················· | ···················· | ···················· | ···················· |
| AB | 1181 | hCoV-19/England/205081168/2020\|EPI_ISL_766292\|2020 | ···················· | ···················· | ···················· | ···················· | ···················· | ···················· |
| AB | 1182 | hCoV-19/England/205081475/2020\|EPI_ISL_766293\|2020 | ···················· | ···················· | ···················· | ···················· | ···················· | ···················· |
| AB | 1183 | hCoV-19/England/205081476/2020\|EPI_ISL_766294\|2020 | ···················· | ···················· | ···················· | ···················· | ···················· | ···················· |
| AB | 1184 | hCoV-19/England/205090056/2020\|EPI_ISL_766295\|2020 | ···················· | ···················· | ···················· | ···················· | ···················· | ···················· |
| AB | 1185 | hCoV-19/England/205090680/2020\|EPI_ISL_766296\|2020 | ···················· | ···················· | ···················· | ···················· | ···················· | ···················· |
| AB | 1186 | hCoV-19/England/205090681/2020\|EPI_ISL_766297\|2020 | ···················· | ·A·················· | ···················· | ···················· | ···················· | ···················· |
| AB | 1187 | hCoV-19/England/205091056/2020\|EPI_ISL_766298\|2020 | ···················· | ···················· | ···················· | ···················· | ···················· | ···················· |
| AB | 1188 | hCoV-19/England/205091067/2020\|EPI_ISL_766299\|2020 | ···················· | ···················· | ···················· | ···················· | ···················· | ···················· |
| AB | 1189 | hCoV-19/USA/NY-Wadsworth-289758-01/2020\|EPI_ISL_76 | ···················· | ·A·················· | ···················· | ···················· | ···················· | ···················· |
| AB | 1190 | hCoV-19/USA/NY-Wadsworth-289759-01/2020\|EPI_ISL_76 | ···················· | ·A·················· | ···················· | ···················· | ···················· | ···················· |
| AB | 1191 | hCoV-19/USA/NY-Wadsworth-289761-01/2020\|EPI_ISL_76 | ···················· | ·A·················· | ···················· | ···················· | ···················· | ···················· |
| AB | 1192 | hCoV-19/USA/NY-Wadsworth-289792-01/2020\|EPI_ISL_76 | ···················· | ···················· | ···················· | ···················· | ···················· | ···················· |
| AB | 1193 | hCoV-19/USA/NY-Wadsworth-289843-01/2020\|EPI_ISL_76 | ···················· | ···················· | ···················· | ···················· | ···················· | ···················· |
| AB | 1194 | hCoV-19/USA/NY-Wadsworth-289845-01/2020\|EPI_ISL_76 | ···················· | ···················· | ···················· | ···················· | ···················· | ···················· |
| AB | 1195 | hCoV-19/USA/NY-Wadsworth-289851-01/2020\|EPI_ISL_76 | ···················· | ···················· | ···················· | ···················· | ···················· | ···················· |
| AB | 1196 | hCoV-19/USA/NY-Wadsworth-289873-01/2020\|EPI_ISL_76 | ···················· | ···················· | ···················· | ···················· | ···················· | ···················· |
| AB | 1197 | hCoV-19/USA/NY-Wadsworth-289895-01/2020\|EPI_ISL_76 | ···················· | ·A·················· | ···················· | ···················· | ···················· | ···················· |
| AB | 1198 | hCoV-19/USA/NY-Wadsworth-289934-01/2020\|EPI_ISL_76 | ···················· | ·············A······ | ···················· | ···················· | ···················· | ···················· |
| AB | 1199 | hCoV-19/USA/NY-Wadsworth-289941-01/2020\|EPI_ISL_76 | ···················· | ···················· | ···················· | ···················· | ···················· | ···················· |
| AB | 1200 | hCoV-19/USA/NY-Wadsworth-289942-01/2020\|EPI_ISL_76 | ···················· | ···················· | ···················· | ···················· | ···················· | ···················· |
| AB | 1201 | hCoV-19/USA/NY-Wadsworth-289945-01/2020\|EPI_ISL_76 | ···················· | ···················· | ···················· | ···················· | ···················· | ···················· |
| AB | 1202 | hCoV-19/USA/NY-Wadsworth-289954-01/2020\|EPI_ISL_76 | ···················· | ···················· | ···················· | ···················· | ···················· | ···················· |
| AB | 1203 | hCoV-19/USA/NY-Wadsworth-289958-01/2020\|EPI_ISL_76 | ···················· | ·A·················· | ···················· | ···················· | ···················· | ···················· |
| AB | 1204 | hCoV-19/USA/NY-Wadsworth-290015-01/2020\|EPI_ISL_76 | ···················· | ···················· | ···················· | ···················· | ···················· | ···················· |
| AB | 1205 | hCoV-19/USA/NY-Wadsworth-290047-01/2020\|EPI_ISL_76 | ···················· | ···················· | ···················· | ···················· | ···················· | ···················· |

| | | | | | | | | |
|---|---|---|---|---|---|---|---|---|
| AB | 1206 | hCoV-19/USA/NY-Wadsworth-290175-01/2020\|EPI_ISL_76 | ···················· | ···················· | ····················· | ···················· | T··················· | ···················· |
| AB | 1207 | hCoV-19/USA/NY-Wadsworth-290270-01/2020\|EPI_ISL_76 | ···················· | ···················· | ····················· | ···················· | ···················· | ···················· |
| AB | 1208 | hCoV-19/USA/NY-Wadsworth-290331-01/2020\|EPI_ISL_76 | ···················· | ···················· | ····················· | ···················· | ···················· | ···················· |
| AB | 1209 | hCoV-19/USA/NY-Wadsworth-290332-01/2020\|EPI_ISL_76 | ···················· | ···················· | ····················· | ···················· | ···················· | ···················· |
| AB | 1210 | hCoV-19/USA/NY-Wadsworth-290473-01/2020\|EPI_ISL_76 | ···················· | ·A·················· | ····················· | ···················· | ···················· | ···················· |
| AB | 1211 | hCoV-19/USA/NY-Wadsworth-290495-01/2020\|EPI_ISL_76 | ···················· | ·A·················· | ····················· | ···················· | ···················· | ···················· |
| AB | 1212 | hCoV-19/USA/NY-Wadsworth-290629-01/2020\|EPI_ISL_76 | ···················· | ·A·················· | ····················· | ···················· | ···················· | ···················· |
| AB | 1213 | hCoV-19/USA/NY-Wadsworth-290630-01/2020\|EPI_ISL_76 | ···················· | ·A·················· | ····················· | ···················· | ···················· | ···················· |
| AB | 1214 | hCoV-19/USA/NY-Wadsworth-290631-01/2020\|EPI_ISL_76 | ···················· | ·A·················· | ····················· | ···················· | ···················· | ···················· |
| AB | 1215 | hCoV-19/USA/NY-Wadsworth-290632-01/2020\|EPI_ISL_76 | ···················· | ·A·················· | ····················· | ···················· | ···················· | ···················· |
| AB | 1216 | hCoV-19/USA/NY-Wadsworth-290633-01/2020\|EPI_ISL_76 | ···················· | ·A·················· | ····················· | ···················· | ···················· | ···················· |
| AB | 1217 | hCoV-19/USA/NY-Wadsworth-290635-01/2020\|EPI_ISL_76 | ···················· | ·A·················· | ····················· | ···················· | ···················· | ···················· |
| AB | 1218 | hCoV-19/USA/NY-Wadsworth-290636-01/2020\|EPI_ISL_76 | ···················· | ·A·················· | ····················· | ···················· | ···················· | ···················· |
| AB | 1219 | hCoV-19/USA/NY-Wadsworth-292264-01/2020\|EPI_ISL_76 | ···················· | ···················· | ····················· | ···················· | ···················· | ···················· |
| AB | 1220 | hCoV-19/USA/NY-Wadsworth-292287-01/2020\|EPI_ISL_76 | ···················· | ·A·················· | ····················· | ···················· | ···················· | ···················· |
| AB | 1221 | hCoV-19/USA/NY-Wadsworth-291670-01/2020\|EPI_ISL_76 | ···················· | ·A·················· | ····················· | ···················· | ···················· | ···················· |
| AB | 1222 | hCoV-19/USA/NY-Wadsworth-291671-01/2020\|EPI_ISL_76 | ···················· | ···················· | ····················· | ···················· | ···················· | ···················· |
| AB | 1223 | hCoV-19/USA/NY-Wadsworth-291674-01/2020\|EPI_ISL_76 | ···················· | ···················· | ····················· | ···················· | ···················· | ···················· |
| AB | 1224 | hCoV-19/USA/NY-Wadsworth-291676-01/2020\|EPI_ISL_76 | ···················· | ···················· | ····················· | ···················· | ···················· | ···················· |
| AB | 1225 | hCoV-19/USA/NY-Wadsworth-291677-01/2020\|EPI_ISL_76 | ···················· | ·A·················· | ····················· | ···················· | ···················· | ···················· |
| AB | 1226 | hCoV-19/USA/NY-Wadsworth-291678-01/2020\|EPI_ISL_76 | ···················· | ·A·················· | ····················· | ···················· | ···················· | ···················· |
| AB | 1227 | hCoV-19/USA/NY-Wadsworth-291679-01/2020\|EPI_ISL_76 | ···················· | ·A·················· | ····················· | ···················· | ···················· | ···················· |
| AB | 1228 | hCoV-19/USA/NY-Wadsworth-291742-01/2020\|EPI_ISL_76 | ···················· | ···················· | ····················· | ···················· | ···················· | ···················· |
| AB | 1229 | hCoV-19/USA/NY-Wadsworth-291743-01/2020\|EPI_ISL_76 | ···················· | ·A·················· | ····················· | ···················· | ···················· | ···················· |
| AB | 1230 | hCoV-19/USA/NY-Wadsworth-291745-01/2020\|EPI_ISL_76 | ···················· | ···················· | ····················· | ···················· | ···················· | ···················· |
| AB | 1231 | hCoV-19/USA/NY-Wadsworth-291747-01/2020\|EPI_ISL_76 | ···················· | ···················· | ····················· | ···················· | ···················· | ···················· |
| AB | 1232 | hCoV-19/USA/NY-Wadsworth-291748-01/2020\|EPI_ISL_76 | ···················· | ···················· | ····················· | ···················· | ···················· | ···················· |
| AB | 1233 | hCoV-19/USA/NY-Wadsworth-291749-01/2020\|EPI_ISL_76 | ···················· | ·A·················· | ····················· | ···················· | ···················· | ···················· |
| AB | 1234 | hCoV-19/USA/NY-Wadsworth-291750-01/2020\|EPI_ISL_76 | ···················· | ·A·················· | ····················· | ···················· | ···················· | ···················· |
| AB | 1235 | hCoV-19/USA/NY-Wadsworth-291752-01/2020\|EPI_ISL_76 | ···················· | ···················· | ····················· | ···················· | ···················· | ···················· |
| AB | 1236 | hCoV-19/USA/NY-Wadsworth-291754-01/2020\|EPI_ISL_76 | ···················· | ·A·················· | ····················· | ···················· | ···················· | ···················· |
| AB | 1237 | hCoV-19/Morocco/FMP-37/2020\|EPI_ISL_768836\|2020-12 | ···················· | ···················· | ····················· | ···················· | ···················· | ···················· |
| AB | 1238 | hCoV-19/Morocco/FMP-38/2020\|EPI_ISL_768837\|2020-12 | ···················· | ···················· | ····················· | ···················· | ···················· | ···················· |
| AB | 1239 | hCoV-19/Morocco/FMP-39/2020\|EPI_ISL_768838\|2020 | ···················· | ···················· | ····················· | ···················· | ···················· | ···················· |
| AB | 1240 | hCoV-19/Morocco/FMP-40/2020\|EPI_ISL_768839\|2020-11 | ···················· | ······A············· | ····················· | ···················· | ···················· | ···················· |
| AB | 1241 | hCoV-19/England/MILK-CAA39A/2020\|EPI_ISL_768849\|20 | ···················· | ···················· | ····················· | ···················· | ···················· | ···················· |
| AB | 1242 | hCoV-19/England/MILK-CAA127/2020\|EPI_ISL_768856\|20 | ···················· | ···················· | ····················· | ···················· | ···················· | ···················· |
| AB | 1243 | hCoV-19/England/MILK-CAA36D/2020\|EPI_ISL_768863\|20 | ···················· | ···················· | ····················· | ···················· | ···················· | ···················· |
| AB | 1244 | hCoV-19/England/CAMC-C72049/2020\|EPI_ISL_768902\|20 | ···················· | ···················· | ····················· | ···················· | ···················· | ···················· |
| AB | 1245 | hCoV-19/England/CAMC-C71E6E/2020\|EPI_ISL_768907\|20 | ···················· | ···················· | ····················· | ···················· | ···················· | ···················· |
| AB | 1246 | hCoV-19/England/CAMC-C7569F/2020\|EPI_ISL_768909\|20 | ···················· | ···················· | ····················· | ···················· | ···················· | ···················· |
| AB | 1247 | hCoV-19/England/CAMC-C72191/2020\|EPI_ISL_768912\|20 | ···················· | ···················· | ····················· | ···················· | ······T············· | ···················· |
| AB | 1248 | hCoV-19/England/CAMC-C74706/2020\|EPI_ISL_768913\|20 | ···················· | ···················· | ····················· | ···················· | ···················· | ···················· |
| AB | 1249 | hCoV-19/England/CAMC-C71E31/2020\|EPI_ISL_768915\|20 | ···················· | ···················· | ····················· | ···················· | ···················· | ···················· |
| AB | 1250 | hCoV-19/England/CAMC-C771AB/2020\|EPI_ISL_768919\|20 | ···················· | ···················· | ····················· | ···················· | ···················· | ···················· |
| AB | 1251 | hCoV-19/England/CAMC-C71D70/2020\|EPI_ISL_768921\|20 | ···················· | ···················· | ····················· | ···················· | ···················· | ···················· |
| AB | 1252 | hCoV-19/England/CAMC-C73716/2020\|EPI_ISL_768929\|20 | ···················· | ···················· | ····················· | ···················· | ···················· | ···················· |
| AB | 1253 | hCoV-19/England/CAMC-C746CD/2020\|EPI_ISL_768930\|20 | ···················· | ···················· | ····················· | ···················· | ···················· | ···················· |
| AB | 1254 | hCoV-19/England/CAMC-C70D53/2020\|EPI_ISL_768932\|20 | ···················· | ···················· | ····················· | ···················· | ···················· | ···················· |
| AB | 1255 | hCoV-19/England/CAMC-C71D25/2020\|EPI_ISL_768933\|20 | ···················· | ···················· | ····················· | ···················· | ···················· | ···················· |
| AB | 1256 | hCoV-19/England/CAMC-C771C9/2020\|EPI_ISL_768937\|20 | ···················· | ···················· | ····················· | ···················· | ···················· | ···················· |
| AB | 1257 | hCoV-19/England/CAMC-C71FB6/2020\|EPI_ISL_768938\|20 | ···················· | ···················· | ····················· | ···················· | ···················· | ···················· |
| AB | 1258 | hCoV-19/England/CAMC-CB8DC8/2020\|EPI_ISL_778231\|20 | ···················· | ···················· | ····················· | ···················· | ···················· | ···················· |
| AB | 1259 | hCoV-19/England/ALDP-CC09AB/2020\|EPI_ISL_778234\|20 | ···················· | ···················· | ····················· | ···················· | ···················· | ···················· |
| AB | 1260 | hCoV-19/England/QEUH-CC2815/2020\|EPI_ISL_778236\|20 | ···················· | ···················· | ····················· | ···················· | ···A················ | ···················· |
| AB | 1261 | hCoV-19/England/ALDP-CC0219/2020\|EPI_ISL_778239\|20 | ···················· | ···················· | ····················· | ···················· | ···················· | ···················· |
| AB | 1262 | hCoV-19/England/CAMC-CB7039/2020\|EPI_ISL_778242\|20 | ···················· | ···················· | ····················· | ···················· | ···················· | ···················· |
| AB | 1263 | hCoV-19/England/QEUH-CC2806/2020\|EPI_ISL_778244\|20 | ···················· | ···················· | ····················· | ···················· | ···················· | ···A················ |
| AB | 1264 | hCoV-19/England/QEUH-CC2E6E/2020\|EPI_ISL_778248\|20 | ···················· | ···················· | ····················· | ···················· | ···················· | ···················· |
| AB | 1265 | hCoV-19/England/CAMC-CB6D15/2020\|EPI_ISL_778255\|20 | ···················· | ···················· | ····················· | ···················· | ···················· | ···················· |
| AB | 1266 | hCoV-19/England/ALDP-CBE15D/2020\|EPI_ISL_778256\|20 | ···················· | ···················· | ···········T········· | ···················· | ···················· | ···················· |
| AB | 1267 | hCoV-19/England/CAMC-CB742E/2020\|EPI_ISL_778257\|20 | ···················· | ···················· | ····················· | ···················· | ···················· | ···················· |
| AB | 1268 | hCoV-19/England/QEUH-CC25F0/2020\|EPI_ISL_778260\|20 | ···················· | ···················· | ····················· | ···················· | ···················· | ···················· |
| AB | 1269 | hCoV-19/England/ALDP-CC09C9/2020\|EPI_ISL_778261\|20 | ···················· | ···················· | ····················· | ···················· | ···················· | ···················· |
| AB | 1270 | hCoV-19/England/QEUH-CC2763/2020\|EPI_ISL_778263\|20 | ···················· | ···················· | ····················· | ···················· | ···················· | ···················· |
| AB | 1271 | hCoV-19/England/QEUH-CC3638/2020\|EPI_ISL_778269\|20 | ···················· | ···················· | ····················· | ···················· | ···················· | ···················· |
| AB | 1272 | hCoV-19/England/CAMC-CB7172/2020\|EPI_ISL_778273\|20 | ···················· | ···················· | ····················· | ···················· | ···················· | ···················· |
| AB | 1273 | hCoV-19/England/QEUH-CC2860/2020\|EPI_ISL_778276\|20 | ···················· | ···················· | ····················· | ···················· | ···················· | ···················· |
| AB | 1274 | hCoV-19/England/QEUH-CC2824/2020\|EPI_ISL_778279\|20 | ···················· | ···················· | ····················· | ···················· | ···A················ | ···················· |
| AB | 1275 | hCoV-19/England/QEUH-CC25B4/2020\|EPI_ISL_778281\|20 | ···················· | ···················· | ····················· | ···················· | ···················· | ···················· |
| AB | 1276 | hCoV-19/England/QEUH-CC3629/2020\|EPI_ISL_778283\|20 | ···················· | ···················· | ····················· | ···················· | ···················· | ···················· |
| AB | 1277 | hCoV-19/England/CAMC-CB6DE8/2020\|EPI_ISL_778285\|20 | ···················· | ···················· | ····················· | ···················· | ···················· | ···················· |
| AB | 1278 | hCoV-19/England/ALDP-CBF4BD/2020\|EPI_ISL_778287\|20 | ···················· | ···················· | ····················· | ···················· | ···················· | ···················· |
| AB | 1279 | hCoV-19/England/CAMC-CB73A9/2020\|EPI_ISL_778291\|20 | ···················· | ···················· | ····················· | ···················· | ···················· | ···················· |
| AB | 1280 | hCoV-19/England/QEUH-C6443F/2020\|EPI_ISL_778297\|20 | ···················· | ···················· | ····················· | ···················· | ···················· | ···················· |
| AB | 1281 | hCoV-19/South Korea/KDCA0373/2020\|EPI_ISL_760157\|2 | ···················· | ···················· | ····················· | ···················· | ···················· | ···················· |
| AB | 1282 | hCoV-19/South Korea/KDCA0374/2020\|EPI_ISL_760158\|2 | ············A······· | ···················· | ····················· | ···················· | ···················· | ···················· |
| AB | 1283 | hCoV-19/South Korea/KDCA0375/2020\|EPI_ISL_760159\|2 | ···················· | ···················· | ····················· | ···················· | ···················· | ···················· |
| AB | 1284 | hCoV-19/South Korea/KDCA0376/2020\|EPI_ISL_760160\|2 | ············A······· | ···················· | ····················· | ···················· | ···················· | ···················· |
| AB | 1285 | hCoV-19/South Korea/KDCA0377/2020\|EPI_ISL_760161\|2 | ···················· | ···················· | ····················· | ···················· | ···················· | ···················· |

| | | | | | | | | |
|---|---|---|---|---|---|---|---|---|
| AB | 1286 | hCoV-19/South Korea/KDCA0378/2020\|EPI_ISL_760162\|2 | .................... | .................... | ..................... | .................... | .................... | .................... |
| AB | 1287 | hCoV-19/South Korea/KDCA0379/2020\|EPI_ISL_760163\|2 | .................... | .................... | ..................... | .................... | .................... | .................... |
| AB | 1288 | hCoV-19/South Korea/KDCA0384/2020\|EPI_ISL_760168\|2 | .................... | .................... | ..................... | .................... | .................... | .................... |
| AB | 1289 | hCoV-19/South Korea/KDCA0385/2020\|EPI_ISL_760169\|2 | .................... | .................... | ..................... | .................... | .................... | .................... |
| AB | 1290 | hCoV-19/South Korea/KDCA0386/2020\|EPI_ISL_760170\|2 | .................... | .................... | ..................... | .................... | .................... | .................... |
| AB | 1291 | hCoV-19/South Korea/KDCA0387/2020\|EPI_ISL_760171\|2 | .................... | .................... | ..................... | .................... | .................... | .................... |
| AB | 1292 | hCoV-19/South Korea/KDCA0388/2020\|EPI_ISL_760172\|2 | .................... | .................... | ..................... | .................... | .................... | .................... |
| AB | 1293 | hCoV-19/South Korea/KDCA0389/2020\|EPI_ISL_760173\|2 | .................... | .................... | ..................... | .................... | .................... | .................... |
| AB | 1294 | hCoV-19/South Korea/KDCA0390/2020\|EPI_ISL_760174\|2 | .................... | .................... | ..................... | .................... | .................... | .................... |
| AB | 1295 | hCoV-19/South Korea/KDCA0391/2020\|EPI_ISL_760175\|2 | .................... | .................... | ..................... | .................... | .................... | .................... |
| AB | 1296 | hCoV-19/South Korea/KDCA0392/2020\|EPI_ISL_760176\|2 | .................... | .................... | ..................... | .................... | .................... | .................... |
| AB | 1297 | hCoV-19/South Korea/KDCA0393/2020\|EPI_ISL_760177\|2 | .................... | .................... | ..................... | .................... | .................... | .................... |
| AB | 1298 | hCoV-19/South Korea/KDCA0394/2020\|EPI_ISL_760178\|2 | .................... | .................... | ..................... | .................... | .................... | .................... |
| AB | 1299 | hCoV-19/South Korea/KDCA0395/2020\|EPI_ISL_760179\|2 | .................... | .................... | ..................... | .................... | .................... | .................... |
| AB | 1300 | hCoV-19/South Korea/KDCA0396/2020\|EPI_ISL_760180\|2 | .................... | .................... | ..................... | .................... | .................... | .................... |
| AB | 1301 | hCoV-19/South Korea/KDCA0397/2020\|EPI_ISL_760181\|2 | .................... | .................... | ..................... | .................... | .................... | .................... |
| AB | 1302 | hCoV-19/South Korea/KDCA0398/2020\|EPI_ISL_760182\|2 | .................... | .................... | ..................... | .................... | .................... | .................... |
| AB | 1303 | hCoV-19/South Korea/KDCA0399/2020\|EPI_ISL_760183\|2 | .................... | .................... | ..................... | .................... | .................... | .................... |
| AB | 1304 | hCoV-19/South Korea/KDCA0400/2020\|EPI_ISL_760184\|2 | .................... | .................... | ..................... | .................... | .................... | .................... |
| AB | 1305 | hCoV-19/South Korea/KDCA0401/2020\|EPI_ISL_760185\|2 | .................... | .................... | ..................... | .................... | .................... | .................... |
| AB | 1306 | hCoV-19/South Korea/KDCA0402/2020\|EPI_ISL_760186\|2 | .................... | .................... | ..................... | .................... | .................... | .................... |
| AB | 1307 | hCoV-19/South Korea/KDCA0403/2020\|EPI_ISL_760187\|2 | .................... | .................... | ..................... | .................... | .................... | .................... |
| AB | 1308 | hCoV-19/South Korea/KDCA0404/2020\|EPI_ISL_760188\|2 | .................... | .................... | ..................... | .................... | .................... | .................... |
| AB | 1309 | hCoV-19/South Korea/KDCA0405/2020\|EPI_ISL_760189\|2 | .................... | .................... | ..................... | .................... | .................... | .................... |
| AB | 1310 | hCoV-19/England/MILK-CA6D72/2020\|EPI_ISL_761493\|20 | .................... | .................... | ..................... | .................... | .................... | .................... |
| AB | 1311 | hCoV-19/Wales/PHWC-4A60F9/2020\|EPI_ISL_765000\|2020 | .................... | .................... | ..................... | .................... | .................... | .................... |
| AB | 1312 | hCoV-19/Wales/PHWC-4A6105/2020\|EPI_ISL_765001\|2020 | .................... | .................... | ..................... | .................... | .................... | .................... |
| AB | 1314 | hCoV-19/Wales/PHWC-4A6132/2020\|EPI_ISL_765003\|2020 | .................... | .................... | ..................... | .................... | .................... | .................... |
| AB | 1315 | hCoV-19/Wales/PHWC-4A6141/2020\|EPI_ISL_765004\|2020 | .................... | .................... | ..................... | .................... | .................... | .................... |
| AB | 1316 | hCoV-19/Wales/PHWC-4A6150/2020\|EPI_ISL_765005\|2020 | .................... | .................... | ..................... | .................... | .................... | .................... |
| AB | 1317 | hCoV-19/Wales/PHWC-4A616F/2020\|EPI_ISL_765006\|2020 | .................... | .................... | ..................... | .................... | .................... | .................... |
| AB | 1318 | hCoV-19/Wales/PHWC-4A617E/2020\|EPI_ISL_765007\|2020 | .................... | .................... | ..................... | .................... | .................... | .................... |
| AB | 1319 | hCoV-19/Wales/PHWC-4A618D/2020\|EPI_ISL_765008\|2020 | .................... | .................... | ..................... | .................... | .................... | .................... |
| AB | 1320 | hCoV-19/Wales/PHWC-4A619C/2020\|EPI_ISL_765009\|2020 | .................... | .................... | ..................... | .................... | .................... | .................... |
| AB | 1321 | hCoV-19/Wales/PHWC-4A61BA/2020\|EPI_ISL_765010\|2020 | .................... | .................... | ..................... | .................... | .................... | .................... |
| AB | 1322 | hCoV-19/Wales/PHWC-4A61C9/2020\|EPI_ISL_765011\|2020 | .................... | .................... | ..................... | .................... | .................... | .................... |
| AB | 1323 | hCoV-19/Wales/PHWC-4A61D8/2020\|EPI_ISL_765012\|2020 | .................... | .................... | ..................... | .................... | .................... | .................... |
| AB | 1324 | hCoV-19/Wales/PHWC-4A6211/2020\|EPI_ISL_765013\|2020 | .................... | .................... | ..................... | .................... | .................... | .................... |
| AB | 1325 | hCoV-19/Wales/PHWC-4A623F/2020\|EPI_ISL_765014\|2020 | .................... | .................... | ..................... | .................... | .................... | .................... |
| AB | 1326 | hCoV-19/Wales/PHWC-4A625D/2020\|EPI_ISL_765015\|2020 | .................... | .................... | ..................... | .................... | .................... | .................... |
| AB | 1327 | hCoV-19/Wales/PHWC-4A626C/2020\|EPI_ISL_765016\|2020 | .................... | .................... | ..................... | .................... | .................... | .................... |
| AB | 1328 | hCoV-19/Wales/PHWC-4A627B/2020\|EPI_ISL_765017\|2020 | .................... | .................... | ..................... | .................... | .................... | .................... |
| AB | 1330 | hCoV-19/Wales/PHWC-4A6299/2020\|EPI_ISL_765019\|2020 | .................... | .................... | ..................... | .................... | .................... | .................... |
| AB | 1331 | hCoV-19/Wales/PHWC-4A62A8/2020\|EPI_ISL_765020\|2020 | .................... | .................... | ..................... | .................... | .................... | .................... |
| AB | 1332 | hCoV-19/Wales/PHWC-4A62C6/2020\|EPI_ISL_765021\|2020 | .................... | .................... | ..................... | .................... | .................... | .................... |
| AB | 1333 | hCoV-19/Wales/PHWC-4A62D5/2020\|EPI_ISL_765022\|2020 | .................... | .................... | ..................... | .................... | .................... | .................... |
| AB | 1334 | hCoV-19/Wales/PHWC-4A62E4/2020\|EPI_ISL_765023\|2020 | .................... | ..........A......... | ..................... | .................... | .................... | .................... |
| AB | 1335 | hCoV-19/Wales/PHWC-4A62F3/2020\|EPI_ISL_765024\|2020 | .................... | .................... | ..................... | .................... | .................... | .................... |
| AB | 1336 | hCoV-19/Wales/PHWC-4A630F/2020\|EPI_ISL_765025\|2020 | .................... | .................... | ..................... | .................... | .................... | .................... |
| AB | 1337 | hCoV-19/Wales/PHWC-4A631E/2020\|EPI_ISL_765026\|2020 | .................... | .................... | ..................... | .................... | .................... | .................... |
| AB | 1338 | hCoV-19/Wales/PHWC-4A633C/2020\|EPI_ISL_765027\|2020 | .................... | .................... | ..................... | .................... | .................... | .................... |
| AB | 1339 | hCoV-19/Wales/PHWC-4A6369/2020\|EPI_ISL_765028\|2020 | .................... | .................... | ..................... | .................... | .................... | .................... |
| AB | 1341 | hCoV-19/Wales/PHWC-4A6396/2020\|EPI_ISL_765030\|2020 | .................... | .................... | ..................... | .................... | .................... | .................... |
| AB | 1342 | hCoV-19/Wales/PHWC-4A63A5/2020\|EPI_ISL_765031\|2020 | .................... | .................... | ..................... | .................... | .................... | .................... |
| AB | 1343 | hCoV-19/Wales/PHWC-4A63C3/2020\|EPI_ISL_765032\|2020 | .................... | .................... | ..................... | .................... | .................... | .................... |
| AB | 1344 | hCoV-19/Wales/PHWC-4A63D2/2020\|EPI_ISL_765033\|2020 | .................... | .................... | ..................... | .................... | .................... | .................... |
| AB | 1345 | hCoV-19/Wales/PHWC-4A63E1/2020\|EPI_ISL_765034\|2020 | .................... | .................... | ..................... | .................... | .................... | .................... |
| AB | 1346 | hCoV-19/Wales/PHWC-4A641B/2020\|EPI_ISL_765035\|2020 | .................... | .................... | ..................... | .................... | .................... | .................... |
| AB | 1347 | hCoV-19/Wales/PHWC-4A642A/2020\|EPI_ISL_765036\|2020 | .................... | .................... | ..................... | .................... | .................... | .................... |
| AB | 1348 | hCoV-19/Wales/PHWC-4A6439/2020\|EPI_ISL_765037\|2020 | .................... | .................... | ..................... | .................... | .................... | .................... |
| AB | 1349 | hCoV-19/Wales/PHWC-4A6448/2020\|EPI_ISL_765038\|2020 | .................... | .................... | ..................... | .................... | .................... | .................... |
| AB | 1350 | hCoV-19/Wales/PHWC-4A6466/2020\|EPI_ISL_765039\|2020 | .................... | .................... | ..................... | .................... | .................... | .................... |
| AB | 1351 | hCoV-19/England/205091160/2020\|EPI_ISL_766300\|2020 | .................... | .................... | ..................... | .................... | .................... | .................... |
| AB | 1352 | hCoV-19/England/205100232/2020\|EPI_ISL_766301\|2020 | .................... | .................... | ..................... | .................... | .................... | .................... |
| AB | 1353 | hCoV-19/England/205100233/2020\|EPI_ISL_766302\|2020 | .................... | .................... | ..................... | .................... | .................... | .................... |
| AB | 1354 | hCoV-19/England/205120383/2020\|EPI_ISL_766303\|2020 | .................... | .................... | ..................... | .................... | .................... | .................... |
| AB | 1355 | hCoV-19/England/205120464/2020\|EPI_ISL_766304\|2020 | .................... | .A.................. | ..................... | .................... | .................... | ..A................. |
| AB | 1356 | hCoV-19/England/205120470/2020\|EPI_ISL_766305\|2020 | .................... | .A.................. | ..................... | .................... | .................... | ..A................. |
| AB | 1357 | hCoV-19/England/205120471/2020\|EPI_ISL_766306\|2020 | .................... | .A.................. | ..................... | .................... | .................... | ..A................. |
| AB | 1358 | hCoV-19/England/205120473/2020\|EPI_ISL_766307\|2020 | .................... | .A.................. | ..................... | .................... | .................... | ..A................. |
| AB | 1359 | hCoV-19/England/205120481/2020\|EPI_ISL_766308\|2020 | .................... | .A.................. | ..................... | .................... | .................... | ..A................. |
| AB | 1360 | hCoV-19/England/205120484/2020\|EPI_ISL_766309\|2020 | .................... | .A.................. | ..................... | .................... | .................... | ..A................. |
| AB | 1361 | hCoV-19/England/205120686/2020\|EPI_ISL_766310\|2020 | .................... | .................... | ..................... | .................... | .................... | .................... |
| AB | 1362 | hCoV-19/England/205120775/2020\|EPI_ISL_766311\|2020 | .................... | .................... | ..................... | .................... | ...T................ | .................... |
| AB | 1363 | hCoV-19/England/205120777/2020\|EPI_ISL_766312\|2020 | .................... | .................... | ..................... | .................... | .................... | .................... |
| AB | 1364 | hCoV-19/England/205120833/2020\|EPI_ISL_766313\|2020 | .................... | .................... | ..................... | .................... | .................... | .................... |
| AB | 1365 | hCoV-19/England/205120834/2020\|EPI_ISL_766314\|2020 | .................... | .................... | ..................... | .................... | .................... | .................... |
| AB | 1366 | hCoV-19/England/205120836/2020\|EPI_ISL_766315\|2020 | .................... | .................... | ..................... | .................... | .................... | .................... |
| AB | 1367 | hCoV-19/England/205120840/2020\|EPI_ISL_766316\|2020 | .................... | .................... | ..................... | .................... | .................... | .................... |
| AB | 1368 | hCoV-19/Gibraltar/205121190/2020\|EPI_ISL_766320\|20 | .................... | .................... | ..................... | .................... | .................... | .................... |

| | | | | | | | | |
|---|---|---|---|---|---|---|---|---|
| AB | 1369 | hCoV-19/Gibraltar/205121191/2020\|EPI_ISL_766321\|20 | ···················· | ···················· | ····················· | ···················· | ···················· | ···················· |
| AB | 1370 | hCoV-19/Gibraltar/205121192/2020\|EPI_ISL_766322\|20 | ···················· | ···················· | ····················· | ···················· | ···················· | ···················· |
| AB | 1371 | hCoV-19/Gibraltar/205121193/2020\|EPI_ISL_766323\|20 | ···················· | ···················· | ····················· | ···················· | ···················· | ···················· |
| AB | 1372 | hCoV-19/Gibraltar/205121194/2020\|EPI_ISL_766324\|20 | ···················· | ···················· | ····················· | ···················· | ···················· | ···················· |
| AB | 1373 | hCoV-19/Gibraltar/205121195/2020\|EPI_ISL_766325\|20 | ···················· | ···················· | ····················· | ···················· | ···T················ | ···················· |
| AB | 1374 | hCoV-19/Gibraltar/205121196/2020\|EPI_ISL_766326\|20 | ···················· | ···················· | ····················· | ···················· | ···················· | ···················· |
| AB | 1375 | hCoV-19/Gibraltar/205121197/2020\|EPI_ISL_766327\|20 | ···················· | ···················· | ····················· | ···················· | ···················· | ···················· |
| AB | 1376 | hCoV-19/Gibraltar/205121198/2020\|EPI_ISL_766328\|20 | ···················· | ···················· | ····················· | ···················· | ···················· | ···················· |
| AB | 1377 | hCoV-19/Gibraltar/205121199/2020\|EPI_ISL_766329\|20 | ···················· | ···················· | ····················· | ···················· | ···················· | ···················· |
| AB | 1378 | hCoV-19/Gibraltar/205121200/2020\|EPI_ISL_766330\|20 | ···················· | ···················· | ····················· | ···················· | ···················· | ···················· |
| AB | 1379 | hCoV-19/Gibraltar/205121202/2020\|EPI_ISL_766331\|20 | ···················· | ···················· | ····················· | ···················· | ···················· | ···················· |
| AB | 1380 | hCoV-19/Gibraltar/205121203/2020\|EPI_ISL_766332\|20 | ···················· | ···················· | ····················· | ···················· | ···················· | ···················· |
| AB | 1381 | hCoV-19/Gibraltar/205121205/2020\|EPI_ISL_766333\|20 | ···················· | ···················· | ····················· | ···················· | ···················· | ···················· |
| AB | 1382 | hCoV-19/Gibraltar/205121206/2020\|EPI_ISL_766334\|20 | ···················· | ···················· | ····················· | ···················· | ···················· | ···················· |
| AB | 1383 | hCoV-19/Gibraltar/205121208/2020\|EPI_ISL_766335\|20 | ···················· | ···················· | ····················· | ···················· | ···················· | ···················· |
| AB | 1384 | hCoV-19/Gibraltar/205121209/2020\|EPI_ISL_766336\|20 | ···················· | ···················· | ····················· | ···················· | ···················· | ···················· |
| AB | 1385 | hCoV-19/Gibraltar/205121210/2020\|EPI_ISL_766337\|20 | ···················· | ···················· | ····················· | ···················· | ···················· | ···················· |
| AB | 1386 | hCoV-19/Gibraltar/205121212/2020\|EPI_ISL_766338\|20 | ···················· | ···················· | ····················· | ···················· | ···················· | ···················· |
| AB | 1387 | hCoV-19/Gibraltar/205121213/2020\|EPI_ISL_766339\|20 | ···················· | ···················· | ····················· | ···················· | ···················· | ···················· |
| AB | 1388 | hCoV-19/Gibraltar/205121214/2020\|EPI_ISL_766340\|20 | ···················· | ·A·················· | ····················· | ···················· | ···················· | ···················· |
| AB | 1389 | hCoV-19/Gibraltar/205121215/2020\|EPI_ISL_766341\|20 | ···················· | ···················· | ····················· | ···················· | ···················· | ···············A···· |
| AB | 1390 | hCoV-19/Gibraltar/205121216/2020\|EPI_ISL_766342\|20 | ···················· | ···················· | ····················· | ···················· | ···················· | ···················· |
| AB | 1391 | hCoV-19/Gibraltar/205121217/2020\|EPI_ISL_766343\|20 | ···················· | ···················· | ····················· | ···················· | ···················· | ···················· |
| AB | 1392 | hCoV-19/Gibraltar/205121218/2020\|EPI_ISL_766344\|20 | ···················· | ···················· | ····················· | ···················· | ···················· | ···················· |
| AB | 1393 | hCoV-19/Gibraltar/205121219/2020\|EPI_ISL_766345\|20 | ···················· | ···················· | ····················· | ···················· | ···················· | ···················· |
| AB | 1394 | hCoV-19/Gibraltar/205121220/2020\|EPI_ISL_766346\|20 | ···················· | ···················· | ····················· | ···················· | ···················· | ···················· |
| AB | 1395 | hCoV-19/Gibraltar/205121221/2020\|EPI_ISL_766347\|20 | ···················· | ···················· | ····················· | ···················· | ···················· | ···················· |
| AB | 1396 | hCoV-19/Gibraltar/205121222/2020\|EPI_ISL_766348\|20 | ···················· | ···················· | ····················· | ···················· | ···················· | ···················· |
| AB | 1397 | hCoV-19/Gibraltar/205121223/2020\|EPI_ISL_766349\|20 | ···················· | ···················· | ····················· | ···················· | ···················· | ···················· |
| AB | 1398 | hCoV-19/Gibraltar/205121224/2020\|EPI_ISL_766350\|20 | ···················· | ···················· | ····················· | ···················· | ···················· | ···················· |
| AB | 1399 | hCoV-19/Gibraltar/205121225/2020\|EPI_ISL_766351\|20 | ···················· | ···················· | ····················· | ···················· | ···················· | ···················· |
| AB | 1400 | hCoV-19/Gibraltar/205121226/2020\|EPI_ISL_766352\|20 | ···················· | ···················· | ····················· | ···················· | ···················· | ···················· |
| AB | 1401 | hCoV-19/Gibraltar/205121227/2020\|EPI_ISL_766353\|20 | ···················· | ···················· | ····················· | ···················· | ···················· | ···················· |
| AB | 1402 | hCoV-19/Gibraltar/205121229/2020\|EPI_ISL_766354\|20 | ···················· | ···················· | ····················· | ···················· | ···················· | ···················· |
| AB | 1403 | hCoV-19/Gibraltar/205121230/2020\|EPI_ISL_766355\|20 | ···················· | ···················· | ····················· | ···················· | ···················· | ···················· |
| AB | 1404 | hCoV-19/Gibraltar/205121231/2020\|EPI_ISL_766356\|20 | ···················· | ···················· | ····················· | ···················· | ···················· | ···················· |
| AB | 1405 | hCoV-19/Gibraltar/205121232/2020\|EPI_ISL_766357\|20 | ···················· | ···················· | ····················· | ···················· | ···················· | ···················· |
| AB | 1406 | hCoV-19/Gibraltar/205121234/2020\|EPI_ISL_766358\|20 | ···················· | ···················· | ····················· | ···················· | ···················· | ···················· |
| AB | 1407 | hCoV-19/Gibraltar/205121235/2020\|EPI_ISL_766359\|20 | ···················· | ···················· | ····················· | ···················· | ···················· | ···················· |
| AB | 1408 | hCoV-19/Gibraltar/205121236/2020\|EPI_ISL_766360\|20 | ···················· | ·A·················· | ····················· | ···················· | ···················· | ···················· |
| AB | 1409 | hCoV-19/Gibraltar/205121237/2020\|EPI_ISL_766361\|20 | ···················· | ···················· | ····················· | ···················· | ···················· | ···················· |
| AB | 1410 | hCoV-19/Gibraltar/205121238/2020\|EPI_ISL_766362\|20 | ···················· | ···················· | ····················· | ···················· | ···················· | ···················· |
| AB | 1411 | hCoV-19/Gibraltar/205121239/2020\|EPI_ISL_766363\|20 | ···················· | ···················· | ····················· | ···················· | ···················· | ···················· |
| AB | 1412 | hCoV-19/Gibraltar/205121242/2020\|EPI_ISL_766364\|20 | ···················· | ···················· | ····················· | ···················· | ···················· | ···················· |
| AB | 1413 | hCoV-19/Gibraltar/205121243/2020\|EPI_ISL_766365\|20 | ···················· | ···················· | ····················· | ···················· | ···················· | ···················· |
| AB | 1414 | hCoV-19/Gibraltar/205121244/2020\|EPI_ISL_766366\|20 | ···················· | ···················· | ····················· | ···················· | ···················· | ···············A···· |
| AB | 1415 | hCoV-19/Gibraltar/205121245/2020\|EPI_ISL_766367\|20 | ···················· | ···················· | ····················· | ···················· | ···················· | ···················· |
| AB | 1416 | hCoV-19/Gibraltar/205121246/2020\|EPI_ISL_766368\|20 | ···················· | ···················· | ····················· | ···················· | ···················· | ···················· |
| AB | 1417 | hCoV-19/Gibraltar/205121247/2020\|EPI_ISL_766369\|20 | ···················· | ···················· | ····················· | ···················· | ···················· | ···················· |
| AB | 1418 | hCoV-19/USA/NY-Wadsworth-291850-01/2020\|EPI_ISL_76 | ···················· | ·A·················· | ····················· | ···················· | ···················· | ···················· |
| AB | 1419 | hCoV-19/USA/NY-Wadsworth-291990-01/2020\|EPI_ISL_76 | ···················· | ···················· | ····················· | ···················· | ···················· | ···················· |
| AB | 1420 | hCoV-19/USA/NY-Wadsworth-292000-01/2020\|EPI_ISL_76 | ···················· | ···················· | ····················· | ···················· | ···················· | ···················· |
| AB | 1421 | hCoV-19/USA/NY-Wadsworth-292002-01/2020\|EPI_ISL_76 | ···················· | ·A·················· | ····················· | ···················· | ···················· | ···················· |
| AB | 1422 | hCoV-19/USA/NY-Wadsworth-292004-01/2020\|EPI_ISL_76 | ···················· | ···················· | ····················· | ···················· | ···················· | ···················· |
| AB | 1423 | hCoV-19/USA/NY-Wadsworth-292054-01/2020\|EPI_ISL_76 | ···················· | ···················· | ····················· | ···················· | ···················· | ···················· |
| AB | 1424 | hCoV-19/USA/NY-Wadsworth-292056-01/2020\|EPI_ISL_76 | ···················· | ···················· | ····················· | ···················· | ···················· | ···················· |
| AB | 1425 | hCoV-19/USA/NY-Wadsworth-292057-01/2020\|EPI_ISL_76 | ···················· | ···················· | ····················· | ···················· | ···················· | ···················· |
| AB | 1426 | hCoV-19/USA/NY-Wadsworth-292061-01/2020\|EPI_ISL_76 | ···················· | ·A·················· | ····················· | ···················· | ···················· | ···················· |
| AB | 1427 | hCoV-19/USA/NY-Wadsworth-292064-01/2020\|EPI_ISL_76 | ···················· | ·A·················· | ····················· | ···················· | ···················· | ···················· |
| AB | 1428 | hCoV-19/USA/NY-Wadsworth-292065-01/2020\|EPI_ISL_76 | ···················· | ···················· | ····················· | ···················· | ···················· | ···················· |
| AB | 1429 | hCoV-19/USA/NY-Wadsworth-291996-01/2020\|EPI_ISL_76 | ···················· | ·A·················· | ····················· | ···················· | ···················· | ···················· |
| AB | 1430 | hCoV-19/USA/NY-Wadsworth-292003-01/2020\|EPI_ISL_76 | ···················· | ···················· | ····················· | ···················· | ···················· | ···················· |
| AB | 1431 | hCoV-19/USA/NY-Wadsworth-292006-01/2020\|EPI_ISL_76 | ···················· | ···················· | ····················· | ···················· | ···················· | ···················· |
| AB | 1432 | hCoV-19/USA/NY-Wadsworth-292227-01/2020\|EPI_ISL_76 | ···················· | ···················· | ····················· | ···················· | ···················· | ···················· |
| AB | 1433 | hCoV-19/USA/NY-Wadsworth-292248-01/2020\|EPI_ISL_76 | ···················· | ···················· | ····················· | ···················· | ···················· | ···················· |
| AB | 1434 | hCoV-19/USA/NY-Wadsworth-292256-01/2020\|EPI_ISL_76 | ···················· | ·A·················· | ····················· | ···················· | ···················· | ···················· |
| AB | 1435 | hCoV-19/USA/NY-Wadsworth-292258-01/2020\|EPI_ISL_76 | ···················· | ···················· | ····················· | ···················· | ···················· | ···················· |
| AB | 1436 | hCoV-19/USA/NY-Wadsworth-292259-01/2020\|EPI_ISL_76 | ···················· | ···················· | ····················· | ···················· | ···················· | ···················· |
| AB | 1437 | hCoV-19/USA/NY-Wadsworth-292263-01/2020\|EPI_ISL_76 | ···················· | ···················· | ····················· | ···················· | ···················· | ···················· |
| AB | 1438 | hCoV-19/USA/NY-Wadsworth-292266-01/2020\|EPI_ISL_76 | ···················· | ·············A······ | ····················· | ···················· | ···················· | ···················· |
| AB | 1439 | hCoV-19/USA/NY-Wadsworth-292271-01/2020\|EPI_ISL_76 | ···················· | ·············A······ | ····················· | ···················· | ···················· | ···················· |
| AB | 1440 | hCoV-19/USA/NY-Wadsworth-292272-01/2020\|EPI_ISL_76 | ···················· | ···················· | ····················· | ···················· | ···················· | ···················· |
| AB | 1441 | hCoV-19/USA/NY-Wadsworth-292273-01/2020\|EPI_ISL_76 | ···················· | ···················· | ····················· | ···················· | ···················· | ···················· |
| AB | 1442 | hCoV-19/USA/NY-Wadsworth-292274-01/2020\|EPI_ISL_76 | ···················· | ···················· | ····················· | ···················· | ···················· | ···················· |
| AB | 1443 | hCoV-19/USA/NY-Wadsworth-292275-01/2020\|EPI_ISL_76 | ···················· | ···················· | ····················· | ···················· | ···················· | ···················· |
| AB | 1444 | hCoV-19/USA/NY-Wadsworth-292278-01/2020\|EPI_ISL_76 | ···················· | ·A·················· | ····················· | ···················· | ···················· | ···················· |
| AB | 1445 | hCoV-19/USA/NY-Wadsworth-292280-01/2020\|EPI_ISL_76 | ···················· | ···················· | ····················· | ···················· | ···················· | ···················· |
| AB | 1446 | hCoV-19/USA/NY-Wadsworth-292286-01/2020\|EPI_ISL_76 | ···················· | ···················· | ····················· | ···················· | ····A··············· | ···················· |
| AB | 1447 | hCoV-19/USA/NY-Wadsworth-292288-01/2020\|EPI_ISL_76 | ···················· | ···················· | ····················· | ···················· | ···················· | ···················· |
| AB | 1448 | hCoV-19/USA/NY-Wadsworth-292289-01/2020\|EPI_ISL_76 | ···················· | ···················· | ····················· | ···················· | ···················· | ···················· |

| | | | | | | | |
|---|---|---|---|---|---|---|---|
| AB | 1449 hCoV-19/USA/NY-Wadsworth-292291-01/2020\|EPI_ISL_76 | ···················· | ···················· | ····················· | ···················· | ···················· | ···················· |
| AB | 1450 hCoV-19/USA/NY-Wadsworth-292296-01/2020\|EPI_ISL_76 | ···················· | ···················· | ····················· | ···················· | ···················· | ···················· |
| AB | 1451 hCoV-19/USA/NY-Wadsworth-292397-01/2020\|EPI_ISL_76 | ···················· | ···················· | ····················· | ···················· | ···················· | ···················· |
| AB | 1452 hCoV-19/USA/NY-Wadsworth-292401-01/2020\|EPI_ISL_76 | ···················· | ···················· | ····················· | ···················· | ···················· | ···················· |
| AB | 1453 hCoV-19/USA/NY-Wadsworth-292413-01/2020\|EPI_ISL_76 | ···················· | ···················· | ····················· | ···················· | ···················· | ···················· |
| AB | 1454 hCoV-19/USA/NY-Wadsworth-292661-01/2020\|EPI_ISL_76 | ···················· | ···················· | ····················· | ···················· | ···················· | ···················· |
| AB | 1455 hCoV-19/USA/NY-Wadsworth-292688-01/2020\|EPI_ISL_76 | ···················· | ···················· | ····················· | ···················· | ···················· | ···················· |
| AB | 1456 hCoV-19/USA/NY-Wadsworth-292444-01/2020\|EPI_ISL_76 | ···················· | ···················· | ····················· | ···················· | ···················· | ···················· |
| AB | 1457 hCoV-19/USA/NY-Wadsworth-292645-01/2020\|EPI_ISL_76 | ···················· | ···················· | ····················· | ···················· | ···················· | ···················· |
| AB | 1458 hCoV-19/USA/NY-Wadsworth-292229-01/2020\|EPI_ISL_76 | ···················· | ·A·················· | ····················· | ···················· | ···················· | ···················· |
| AB | 1459 hCoV-19/USA/NY-Wadsworth-292230-01/2020\|EPI_ISL_76 | ···················· | ·A·················· | ····················· | ···················· | ···················· | ···················· |
| AB | 1460 hCoV-19/USA/NY-Wadsworth-292231-01/2020\|EPI_ISL_76 | ···················· | ·A·················· | ····················· | ···················· | ···················· | ···················· |
| AB | 1461 hCoV-19/USA/NY-Wadsworth-292233-01/2020\|EPI_ISL_76 | ···················· | ·A·················· | ····················· | ···················· | ···················· | ···················· |
| AB | 1462 hCoV-19/USA/NY-Wadsworth-292234-01/2020\|EPI_ISL_76 | ···················· | ···················· | ····················· | ···················· | ···················· | ···················· |
| AB | 1463 hCoV-19/USA/NY-Wadsworth-292238-01/2020\|EPI_ISL_76 | ···················· | ···················· | ····················· | ···················· | ···················· | ···················· |
| AB | 1464 hCoV-19/USA/NY-Wadsworth-292372-01/2020\|EPI_ISL_76 | ···················· | ···················· | ····················· | ···················· | ···················· | ···················· |
| AB | 1465 hCoV-19/USA/NY-Wadsworth-292376-01/2020\|EPI_ISL_76 | ···················· | ···················· | ····················· | ···················· | ···················· | ···················· |
| AB | 1466 hCoV-19/USA/NY-Wadsworth-292380-01/2020\|EPI_ISL_76 | ···················· | ·A·················· | ····················· | ···················· | ···················· | ···················· |
| AB | 1467 hCoV-19/USA/NY-Wadsworth-292381-01/2020\|EPI_ISL_76 | ···················· | ···················· | ····················· | ···················· | ···················· | ···················· |
| AB | 1468 hCoV-19/USA/NY-Wadsworth-292382-01/2020\|EPI_ISL_76 | ···················· | ·A·················· | ····················· | ···················· | ···················· | ···················· |
| AB | 1469 hCoV-19/USA/NY-Wadsworth-292383-01/2020\|EPI_ISL_76 | ···················· | ·A·················· | ····················· | ···················· | ···················· | ···················· |
| AB | 1470 hCoV-19/USA/NY-Wadsworth-292385-01/2020\|EPI_ISL_76 | ···················· | ···················· | ····················· | ···················· | ···················· | ···················· |
| AB | 1471 hCoV-19/USA/NY-Wadsworth-292386-01/2020\|EPI_ISL_76 | ···················· | ···················· | ····················· | ···················· | ···················· | ···················· |
| AB | 1472 hCoV-19/USA/NY-Wadsworth-292388-01/2020\|EPI_ISL_76 | ···················· | ···················· | ····················· | ···················· | ···················· | ···················· |
| AB | 1473 hCoV-19/USA/NY-Wadsworth-292394-01/2020\|EPI_ISL_76 | ···················· | ···················· | ····················· | ···················· | ···················· | ···················· |
| AB | 1474 hCoV-19/USA/NY-Wadsworth-292395-01/2020\|EPI_ISL_76 | ···················· | ···················· | ····················· | ···················· | ···················· | ···················· |
| AB | 1475 hCoV-19/USA/NY-Wadsworth-292398-01/2020\|EPI_ISL_76 | ···················· | ·A·················· | ····················· | ···················· | ···················· | ···················· |
| AB | 1476 hCoV-19/USA/NY-Wadsworth-292400-01/2020\|EPI_ISL_76 | ···················· | ···················· | ····················· | ···················· | ···················· | ···················· |
| AB | 1477 hCoV-19/USA/NY-Wadsworth-292411-01/2020\|EPI_ISL_76 | ···················· | ···················· | ····················· | ···················· | ···················· | ···················· |
| AB | 1478 hCoV-19/USA/NY-Wadsworth-292414-01/2020\|EPI_ISL_76 | ···················· | ···················· | ····················· | ···················· | ···················· | ···················· |
| AB | 1479 hCoV-19/USA/NY-Wadsworth-292415-01/2020\|EPI_ISL_76 | ···················· | ···················· | ····················· | ···················· | ···················· | ···················· |
| AB | 1480 hCoV-19/USA/NY-Wadsworth-292442-01/2020\|EPI_ISL_76 | ···················· | ·············M······ | ····················· | ···················· | ···················· | ···················· |
| AB | 1481 hCoV-19/USA/NY-Wadsworth-292443-01/2020\|EPI_ISL_76 | ···················· | ···················· | ····················· | ···················· | ···················· | ···················· |
| AB | 1482 hCoV-19/USA/NY-Wadsworth-292445-01/2020\|EPI_ISL_76 | ···················· | ···················· | ····················· | ···················· | ···················· | ···················· |
| AB | 1483 hCoV-19/USA/NY-Wadsworth-292446-01/2020\|EPI_ISL_76 | ···················· | ···················· | ····················· | ···················· | ···················· | ···················· |
| AB | 1484 hCoV-19/USA/NY-Wadsworth-292447-01/2020\|EPI_ISL_76 | ···················· | ···················· | ····················· | ···················· | ···················· | ···················· |
| AB | 1485 hCoV-19/USA/NY-Wadsworth-292451-01/2020\|EPI_ISL_76 | ···················· | ···················· | ····················· | ···················· | ····A··············· | ···················· |
| AB | 1486 hCoV-19/USA/NY-Wadsworth-292470-01/2020\|EPI_ISL_76 | ···················· | ···················· | ····················· | ···················· | ···················· | ···················· |
| AB | 1487 hCoV-19/USA/NY-Wadsworth-292474-01/2020\|EPI_ISL_76 | ···················· | ···················· | ····················· | ···················· | ····A··············· | ···················· |
| AB | 1488 hCoV-19/USA/NY-Wadsworth-292643-01/2020\|EPI_ISL_76 | ···················· | ···················· | ····················· | ···················· | ···················· | ···················· |
| AB | 1489 hCoV-19/USA/NY-Wadsworth-292644-01/2020\|EPI_ISL_76 | ···················· | ···················· | ····················· | ···················· | ···················· | ···················· |
| AB | 1490 hCoV-19/USA/NY-Wadsworth-292646-01/2020\|EPI_ISL_76 | ···················· | ···················· | ····················· | ···················· | ···················· | ···················· |
| AB | 1491 hCoV-19/USA/NY-Wadsworth-292647-01/2020\|EPI_ISL_76 | ···················· | ·A·················· | ····················· | ···················· | ···················· | ···················· |
| AB | 1492 hCoV-19/USA/NY-Wadsworth-292648-01/2020\|EPI_ISL_76 | ···················· | ···················· | ····················· | ···················· | ···················· | ···················· |
| AB | 1493 hCoV-19/USA/NY-Wadsworth-292649-01/2020\|EPI_ISL_76 | ···················· | ·A·················· | ····················· | ···················· | ···················· | ···················· |
| AB | 1494 hCoV-19/USA/NY-Wadsworth-292651-01/2020\|EPI_ISL_76 | ···················· | ···················· | ····················· | ···················· | ···················· | ···················· |
| AB | 1495 hCoV-19/USA/NY-Wadsworth-292653-01/2020\|EPI_ISL_76 | ···················· | ·A·················· | ····················· | ···················· | ···················· | ···················· |
| AB | 1496 hCoV-19/USA/NY-Wadsworth-292655-01/2020\|EPI_ISL_76 | ···················· | ···················· | ····················· | ···················· | ···················· | ···················· |
| AB | 1497 hCoV-19/USA/NY-Wadsworth-292656-01/2020\|EPI_ISL_76 | ···················· | ·A·················· | ····················· | ···················· | ···················· | ···················· |
| AB | 1498 hCoV-19/USA/NY-Wadsworth-292657-01/2020\|EPI_ISL_76 | ···················· | ···················· | ····················· | ···················· | ···················· | ···················· |
| AB | 1499 hCoV-19/England/ALDP-C8A178/2020\|EPI_ISL_768900\|20 | ···················· | ·A·················· | ····················· | ···················· | ···················· | ···················· |
| AB | 1500 hCoV-19/England/CAMC-C70B59/2020\|EPI_ISL_768948\|20 | ···················· | ···················· | ····················· | ···················· | ···················· | ···················· |
| AB | 1501 hCoV-19/England/CAMC-C76731/2020\|EPI_ISL_768949\|20 | ···················· | ···················· | ····················· | ···················· | ···················· | ···················· |
| AB | 1502 hCoV-19/England/CAMC-C7488B/2020\|EPI_ISL_768950\|20 | ···················· | ···················· | ····················· | ···················· | ···················· | ···················· |
| AB | 1503 hCoV-19/England/CAMC-C70CA1/2020\|EPI_ISL_768952\|20 | ···················· | ···················· | ····················· | ···················· | ···················· | ···················· |
| AB | 1504 hCoV-19/England/CAMC-C76704/2020\|EPI_ISL_768953\|20 | ···················· | ···················· | ····················· | ···················· | ···················· | ···················· |
| AB | 1505 hCoV-19/England/CAMC-C70C74/2020\|EPI_ISL_768959\|20 | ···················· | ···················· | ····················· | ···················· | ···················· | ···················· |
| AB | 1506 hCoV-19/England/CAMC-C71E40/2020\|EPI_ISL_768970\|20 | ···················· | ···················· | ····················· | ···················· | ···················· | ···················· |
| AB | 1507 hCoV-19/Wales/CAMC-C755CF/2020\|EPI_ISL_768971\|2020 | ···················· | ···················· | ····················· | ···················· | ···················· | ···················· |
| AB | 1508 hCoV-19/England/CAMC-C72173/2020\|EPI_ISL_768973\|20 | ···················· | ···················· | ····················· | ···················· | ···················· | ···················· |
| AB | 1509 hCoV-19/England/CAMC-C71C0A/2020\|EPI_ISL_768977\|20 | ···················· | ······A············· | ····················· | ···················· | ···················· | ···················· |
| AB | 1510 hCoV-19/England/CAMC-C7487C/2020\|EPI_ISL_768980\|20 | ···················· | ···················· | ····················· | ···················· | ···················· | ···················· |
| AB | 1511 hCoV-19/England/CAMC-C71D16/2020\|EPI_ISL_768981\|20 | ···················· | ···················· | ····················· | ···················· | ···················· | ···················· |
| AB | 1512 hCoV-19/England/CAMC-C74636/2020\|EPI_ISL_768984\|20 | ···················· | ···················· | ····················· | ···················· | ···················· | ···················· |
| AB | 1513 hCoV-19/England/CAMC-C749A6/2020\|EPI_ISL_768987\|20 | ···················· | ···················· | ····················· | ···················· | ···················· | ···················· |
| AB | 1514 hCoV-19/Italy/CAM-TIGEM-372/2020\|EPI_ISL_777000\|20 | ···················· | ···················· | ····················· | ···················· | ······T············· | ···················· |
| AB | 1515 hCoV-19/Italy/CAM-TIGEM-378/2020\|EPI_ISL_777001\|20 | ···················· | ···················· | ····················· | ···················· | ······T············· | ···················· |
| AB | 1516 hCoV-19/Italy/CAM-TIGEM-382/2020\|EPI_ISL_777003\|20 | ···················· | ···················· | ····················· | ···················· | ···················· | ···················· |
| AB | 1517 hCoV-19/Italy/CAM-TIGEM-370/2020\|EPI_ISL_777004\|20 | ···················· | ···················· | ····················· | ···················· | ···················· | ···················· |
| AB | 1518 hCoV-19/Italy/CAM-TIGEM-369/2020\|EPI_ISL_777005\|20 | ···················· | ···················· | ····················· | ···················· | ···················· | ···················· |
| AB | 1519 hCoV-19/Italy/CAM-TIGEM-376/2020\|EPI_ISL_777006\|20 | ···················· | ···················· | ····················· | ···················· | ···················· | ···················· |
| AB | 1520 hCoV-19/Italy/CAM-TIGEM-383/2020\|EPI_ISL_777007\|20 | ···················· | ···················· | ····················· | ···················· | ···················· | ···················· |
| AB | 1521 hCoV-19/Italy/CAM-TIGEM-308/2020\|EPI_ISL_777008\|20 | ···················· | ···················· | ····················· | ···················· | ······T············· | ···················· |
| AB | 1522 hCoV-19/Italy/CAM-TIGEM-323/2020\|EPI_ISL_777009\|20 | ···················· | ···················· | ····················· | ···················· | ······T············· | ···················· |
| AB | 1523 hCoV-19/England/QEUH-C8F5B3/2020\|EPI_ISL_777013\|20 | ···················· | ···················· | ····················· | ···················· | ···················· | ···················· |
| AB | 1524 hCoV-19/England/ALDP-C8C000/2020\|EPI_ISL_777018\|20 | ···················· | ···················· | ····················· | ···················· | ···················· | ···················· |
| AB | 1525 hCoV-19/England/MILK-C6F6B6/2020\|EPI_ISL_778304\|20 | ···················· | ···················· | ····················· | ···················· | ···················· | ···················· |
| AB | 1526 hCoV-19/England/MILK-C6F513/2020\|EPI_ISL_778308\|20 | ···················· | ···················· | ····················· | ···················· | ···················· | ···················· |
| AB | 1527 hCoV-19/England/MILK-C6F5B9/2020\|EPI_ISL_778312\|20 | ···················· | ···················· | ····················· | ···················· | ···················· | ···················· |
| AB | 1528 hCoV-19/England/ALDP-C6B9C0/2020\|EPI_ISL_778315\|20 | ···················· | ···················· | ····················· | ···················· | ···················· | ···················· |

| | | | | | | | |
|---|---|---|---|---|---|---|---|
| AB | 1529 hCoV-19/England/MILK-C6F4BC/2020\|EPI_ISL_778316\|20 | ···················· | ···················· | ···················· | ···················· | ······T············· | ···················· |
| AB | 1530 hCoV-19/England/MILK-C6F540/2020\|EPI_ISL_778317\|20 | ···················· | ···················· | ···················· | ···················· | ···················· | ···················· |
| AB | 1531 hCoV-19/England/MILK-C6F6A7/2020\|EPI_ISL_778319\|20 | ···················· | ···················· | ···················· | ···················· | ···················· | ···················· |
| AB | 1532 hCoV-19/England/QEUH-C643C9/2020\|EPI_ISL_778322\|20 | ···················· | ···················· | ···················· | ···················· | ···················· | ···················· |
| AB | 1533 hCoV-19/England/QEUH-C65331/2020\|EPI_ISL_778324\|20 | ···················· | ···················· | ···················· | ···················· | ······T············· | ···················· |
| AB | 1534 hCoV-19/England/ALDP-C6A16F/2020\|EPI_ISL_778328\|20 | ···················· | ···················· | ···················· | ···················· | ···················· | ···················· |
| AB | 1535 hCoV-19/England/QEUH-C654E3/2020\|EPI_ISL_778332\|20 | ···················· | ···················· | ···················· | ···················· | ···················· | ···················· |
| AB | 1536 hCoV-19/England/ALDP-C6BEEF/2020\|EPI_ISL_778333\|20 | ···················· | ···················· | ···················· | ···················· | ···················· | ···················· |
| AB | 1537 hCoV-19/South Korea/KDCA0416/2020\|EPI_ISL_760200\|2 | ···················· | ···················· | ···················· | ···················· | ···················· | ···················· |
| AB | 1538 hCoV-19/South Korea/KDCA0417/2020\|EPI_ISL_760201\|2 | ···················· | ···················· | ···················· | ···················· | ···················· | ···················· |
| AB | 1539 hCoV-19/South Korea/KDCA0419/2020\|EPI_ISL_760203\|2 | ···················· | ···················· | ···················· | ···················· | ···················· | ···················· |
| AB | 1540 hCoV-19/South Korea/KDCA0420/2020\|EPI_ISL_760204\|2 | ···················· | ···················· | ···················· | ···················· | ······T············· | ···················· |
| AB | 1541 hCoV-19/South Korea/KDCA0421/2020\|EPI_ISL_760205\|2 | ···················· | ···················· | ···················· | ···················· | ···················· | ···················· |
| AB | 1542 hCoV-19/South Korea/KDCA0422/2020\|EPI_ISL_760206\|2 | ···················· | ···················· | ···················· | ···················· | ···················· | ···················· |
| AB | 1543 hCoV-19/South Korea/KDCA0423/2020\|EPI_ISL_760207\|2 | ···················· | ···················· | ···················· | ···················· | ···················· | ···················· |
| AB | 1544 hCoV-19/South Korea/KDCA0424/2020\|EPI_ISL_760208\|2 | ···················· | ···················· | ···················· | ···················· | ···················· | ···················· |
| AB | 1545 hCoV-19/South Korea/KDCA0425/2020\|EPI_ISL_760209\|2 | ···················· | ···················· | ···················· | ···················· | ···················· | ···················· |
| AB | 1546 hCoV-19/South Korea/KDCA0426/2020\|EPI_ISL_760210\|2 | ···················· | ···················· | ···················· | ···················· | ···················· | ···················· |
| AB | 1547 hCoV-19/South Korea/KDCA0427/2020\|EPI_ISL_760211\|2 | ···················· | ···················· | ···················· | ···················· | ···················· | ···················· |
| AB | 1548 hCoV-19/South Korea/KDCA0428/2020\|EPI_ISL_760212\|2 | ···················· | ···················· | ···················· | ···················· | ···················· | ···················· |
| AB | 1549 hCoV-19/South Korea/KDCA0432/2020\|EPI_ISL_760216\|2 | ···················· | ···················· | ···················· | ···················· | ···················· | ···················· |
| AB | 1550 hCoV-19/South Korea/KDCA0433/2020\|EPI_ISL_760217\|2 | ···················· | ···················· | ···················· | ···················· | ···················· | ···················· |
| AB | 1551 hCoV-19/South Korea/KDCA0434/2020\|EPI_ISL_760218\|2 | ···················· | ···················· | ···················· | ···················· | ···················· | ···················· |
| AB | 1552 hCoV-19/South Korea/KDCA0435/2020\|EPI_ISL_760219\|2 | ···················· | ···················· | ···················· | ···················· | ···················· | ···················· |
| AB | 1553 hCoV-19/South Korea/KDCA0436/2020\|EPI_ISL_760220\|2 | ···················· | ···················· | ···················· | ···················· | ···················· | ···················· |
| AB | 1554 hCoV-19/South Korea/KDCA0437/2020\|EPI_ISL_760221\|2 | ···················· | ···················· | ···················· | ···················· | ···················· | ···················· |
| AB | 1555 hCoV-19/South Korea/KDCA0438/2020\|EPI_ISL_760222\|2 | ···················· | ···················· | ···················· | ···················· | ···················· | ···················· |
| AB | 1556 hCoV-19/South Korea/KDCA0439/2020\|EPI_ISL_760223\|2 | ···················· | ···················· | ···················· | ···················· | ···················· | ···················· |
| AB | 1557 hCoV-19/South Korea/KDCA0440/2020\|EPI_ISL_760224\|2 | ···················· | ···················· | ···················· | ···················· | ···················· | ···················· |
| AB | 1558 hCoV-19/South Korea/KDCA0441/2020\|EPI_ISL_760225\|2 | ···················· | ···················· | ···················· | ···················· | ···················· | ···················· |
| AB | 1559 hCoV-19/South Korea/KDCA0442/2020\|EPI_ISL_760226\|2 | ···················· | ·A·················· | ···················· | ···················· | ···················· | ···················· |
| AB | 1560 hCoV-19/South Korea/KDCA0443/2020\|EPI_ISL_760227\|2 | ···················· | ···················· | ···················· | ···················· | ···················· | ···················· |
| AB | 1561 hCoV-19/South Korea/KDCA0444/2020\|EPI_ISL_760228\|2 | ···················· | ···················· | ···················· | ···················· | ···················· | ···················· |
| AB | 1562 hCoV-19/South Korea/KDCA0445/2020\|EPI_ISL_760229\|2 | ···················· | ···················· | ···················· | ···················· | ···················· | ···················· |
| AB | 1563 hCoV-19/South Korea/KDCA0446/2020\|EPI_ISL_760230\|2 | ···················· | ···················· | ···················· | ···················· | ···················· | ···················· |
| AB | 1564 hCoV-19/South Korea/KDCA0448/2020\|EPI_ISL_760232\|2 | ···················· | ···················· | ···················· | ···················· | ···················· | ···················· |
| AB | 1565 hCoV-19/South Korea/KDCA0449/2020\|EPI_ISL_760233\|2 | ···················· | ···················· | ···················· | ···················· | ···················· | ···················· |
| AB | 1566 hCoV-19/South Korea/KDCA0450/2020\|EPI_ISL_760234\|2 | ···················· | ···················· | ···················· | ···················· | ···················· | ···················· |
| AB | 1567 hCoV-19/South Korea/KDCA0451/2020\|EPI_ISL_760235\|2 | ···················· | ···················· | ···················· | ···················· | ···················· | ···················· |
| AB | 1568 hCoV-19/South Korea/KDCA0452/2020\|EPI_ISL_760236\|2 | ···················· | ···················· | ···················· | ···················· | ···················· | ···················· |
| AB | 1569 hCoV-19/South Korea/KDCA0453/2020\|EPI_ISL_760237\|2 | ···················· | ···················· | ···················· | ···················· | ···················· | ···················· |
| AB | 1570 hCoV-19/South Korea/KDCA0454/2020\|EPI_ISL_760238\|2 | ···················· | ···················· | ···················· | ···················· | ···················· | ···················· |
| AB | 1571 hCoV-19/South Korea/KDCA0455/2020\|EPI_ISL_760239\|2 | ···················· | ···················· | ···················· | ···················· | ···················· | ···················· |
| AB | 1572 hCoV-19/South Korea/KDCA0456/2020\|EPI_ISL_760240\|2 | ···················· | ···················· | ···················· | ···················· | ···················· | ···················· |
| AB | 1573 hCoV-19/South Korea/KDCA0457/2020\|EPI_ISL_760241\|2 | ···················· | ···················· | ···················· | ···················· | ···················· | ···················· |
| AB | 1574 hCoV-19/South Korea/KDCA0458/2020\|EPI_ISL_760242\|2 | ···················· | ···················· | ···················· | ···················· | ···················· | ···················· |
| AB | 1575 hCoV-19/South Korea/KDCA0459/2020\|EPI_ISL_760243\|2 | ···················· | ···················· | ···················· | ···················· | ···················· | ···················· |
| AB | 1576 hCoV-19/South Korea/KDCA0460/2020\|EPI_ISL_760244\|2 | ···················· | ···················· | ···················· | ···················· | ···················· | ···················· |
| AB | 1577 hCoV-19/South Korea/KDCA0461/2020\|EPI_ISL_760245\|2 | ···················· | ···················· | ···················· | ···················· | ···················· | ···················· |
| AB | 1578 hCoV-19/England/QEUH-CC5DF5/2020\|EPI_ISL_760251\|20 | ···················· | ···················· | ···················· | ···················· | ···················· | ···················· |
| AB | 1579 hCoV-19/England/QEUH-CC2BD0/2020\|EPI_ISL_760252\|20 | ···················· | ···················· | ···················· | ···················· | ···················· | ···················· |
| AB | 1580 hCoV-19/England/QEUH-CC2684/2020\|EPI_ISL_760255\|20 | ···················· | ···················· | ···················· | ···················· | ···················· | ···················· |
| AB | 1581 hCoV-19/England/QEUH-CC2323/2020\|EPI_ISL_760257\|20 | ···················· | ···················· | ···················· | ···················· | ···················· | ···················· |
| AB | 1582 hCoV-19/England/QEUH-CC5B91/2020\|EPI_ISL_760258\|20 | ···················· | ···················· | ···················· | ···················· | ······T············· | ···················· |
| AB | 1583 hCoV-19/England/QEUH-CC596A/2020\|EPI_ISL_760260\|20 | ···················· | ···················· | ···················· | ···················· | ···················· | ···················· |
| AB | 1584 hCoV-19/England/ALDP-CBE1F3/2020\|EPI_ISL_760261\|20 | ···················· | ···················· | ···················· | ···················· | ···················· | ···················· |
| AB | 1585 hCoV-19/England/QEUH-CC7ADE/2020\|EPI_ISL_760262\|20 | ···················· | ···················· | ···················· | ···················· | ···················· | ···················· |
| AB | 1586 hCoV-19/England/ALDP-CBFE46/2020\|EPI_ISL_760266\|20 | ···················· | ···················· | ···················· | ···················· | ···················· | ···················· |
| AB | 1587 hCoV-19/England/MILK-CAA01B/2020\|EPI_ISL_761517\|20 | ···················· | ···················· | ···················· | ···················· | ···················· | ···················· |
| AB | 1588 hCoV-19/England/MILK-CA7139/2020\|EPI_ISL_761518\|20 | ···················· | ···················· | ···················· | ···················· | ···················· | ···················· |
| AB | 1589 hCoV-19/England/MILK-CA6B87/2020\|EPI_ISL_761526\|20 | ···················· | ···················· | ···················· | ···················· | ···················· | ···················· |
| AB | 1590 hCoV-19/England/MILK-CA6A20/2020\|EPI_ISL_761527\|20 | ···················· | ···················· | ···················· | ···················· | ···················· | ···················· |
| AB | 1591 hCoV-19/England/MILK-CA6A11/2020\|EPI_ISL_761529\|20 | ···················· | ···················· | ···················· | ···················· | ···················· | ···················· |
| AB | 1592 hCoV-19/England/MILK-CA6B96/2020\|EPI_ISL_761547\|20 | ···················· | ···················· | ···················· | ···················· | ···················· | ···················· |
| AB | 1593 hCoV-19/England/MILK-CA9FA6/2020\|EPI_ISL_761551\|20 | ···················· | ···················· | ···················· | ···················· | ···················· | ···················· |
| AB | 1594 hCoV-19/England/MILK-CA6DCD/2020\|EPI_ISL_761567\|20 | ···················· | ···················· | ···················· | ···················· | ···················· | ···················· |
| AB | 1595 hCoV-19/England/MILK-CA6808/2020\|EPI_ISL_761569\|20 | ···················· | ···················· | ···················· | ···················· | ······T············· | ···················· |
| AB | 1596 hCoV-19/England/MILK-CA646E/2020\|EPI_ISL_761582\|20 | ···················· | ···················· | ···················· | ···················· | ···················· | ···················· |
| AB | 1597 hCoV-19/England/MILK-CA6817/2020\|EPI_ISL_761583\|20 | ···················· | ···················· | ···················· | ···················· | ······T············· | ···················· |
| AB | 1598 hCoV-19/England/MILK-CA64C8/2020\|EPI_ISL_761585\|20 | ···················· | ···················· | ···················· | ···················· | ···················· | ···················· |
| AB | 1599 hCoV-19/England/MILK-CA9FD3/2020\|EPI_ISL_761587\|20 | ···················· | ···················· | ···················· | ···················· | ···················· | ···················· |
| AB | 1600 hCoV-19/England/MILK-CA6D27/2020\|EPI_ISL_761591\|20 | ···················· | ···················· | ···················· | ···················· | ···················· | ···················· |
| AB | 1601 hCoV-19/England/QEUH-C7FEBB/2020\|EPI_ISL_762801\|20 | ···················· | ···················· | ···················· | ···················· | ···················· | ···················· |
| AB | 1602 hCoV-19/England/CAMC-C804D3/2020\|EPI_ISL_762806\|20 | ···················· | ···················· | ···················· | ···················· | ···················· | ···················· |
| AB | 1603 hCoV-19/England/CAMC-C824C2/2020\|EPI_ISL_762807\|20 | ···················· | ···················· | ···················· | ···················· | ···················· | ···················· |
| AB | 1604 hCoV-19/England/CAMC-C81E4D/2020\|EPI_ISL_762825\|20 | ···················· | ·A·················· | ···················· | ···················· | ···················· | ··A················· |
| AB | 1605 hCoV-19/England/CAMC-C82F85/2020\|EPI_ISL_762827\|20 | ···················· | ···················· | ···················· | ···················· | ···················· | ···················· |
| AB | 1606 hCoV-19/England/ALDP-C886F3/2020\|EPI_ISL_762832\|20 | ···················· | ···················· | ··········T········· | ···················· | ···················· | ···················· |
| AB | 1607 hCoV-19/England/CAMC-C83731/2020\|EPI_ISL_762834\|20 | ···················· | ···················· | ···················· | ···················· | ···················· | ···················· |
| AB | 1608 hCoV-19/England/ALDP-C889FA/2020\|EPI_ISL_762839\|20 | ···················· | ···················· | ···················· | ···················· | ···················· | ···················· |

| | | | | | | | | |
|---|---|---|---|---|---|---|---|---|
| AB | 1609 | hCoV-19/England/CAMC-C838E3/2020\|EPI_ISL_762840\|20 | ···················· | ···················· | ···················· | ···················· | ···················· | ···················· |
| AB | 1610 | hCoV-19/England/ALDP-C889AF/2020\|EPI_ISL_762841\|20 | ···················· | ·A·················· | ···················· | ···················· | ···················· | ···A················ |
| AB | 1611 | hCoV-19/England/CAMC-C824D1/2020\|EPI_ISL_762842\|20 | ···················· | ···················· | ···················· | ···················· | ···················· | ···················· |
| AB | 1612 | hCoV-19/England/ALDP-C883B0/2020\|EPI_ISL_762843\|20 | ···················· | ···················· | ···················· | ···················· | ···················· | ···················· |
| AB | 1613 | hCoV-19/England/ALDP-C8A10F/2020\|EPI_ISL_762847\|20 | ···················· | ···················· | ···················· | ···················· | ···················· | ···················· |
| AB | 1614 | hCoV-19/England/QEUH-C7F862/2020\|EPI_ISL_762861\|20 | ···················· | ···················· | ···················· | ···················· | ···················· | ···················· |
| AB | 1615 | hCoV-19/England/MILK-C7AE83/2020\|EPI_ISL_762864\|20 | ···················· | ···················· | ···················· | ···················· | ···················· | ···················· |
| AB | 1616 | hCoV-19/England/QEUH-C7F871/2020\|EPI_ISL_762866\|20 | ···················· | ···················· | ···················· | ···················· | ···················· | ···················· |
| AB | 1617 | hCoV-19/England/MILK-C7ABB9/2020\|EPI_ISL_762867\|20 | ···················· | ···················· | ···················· | ···················· | ···················· | ···················· |
| AB | 1618 | hCoV-19/England/ALDP-C884AE/2020\|EPI_ISL_762869\|20 | ···················· | ···················· | ···················· | ···················· | ······T············· | ···················· |
| AB | 1619 | hCoV-19/England/ALDP-C89285/2020\|EPI_ISL_762878\|20 | ···················· | ···················· | ···················· | ···················· | ···················· | ···················· |
| AB | 1620 | hCoV-19/England/ALDP-C8B283/2020\|EPI_ISL_762883\|20 | ···················· | ···················· | ···················· | ···················· | ···················· | ···················· |
| AB | 1621 | hCoV-19/England/QEUH-C7D49D/2020\|EPI_ISL_762888\|20 | ···················· | ···················· | ···················· | ···················· | ···················· | ···················· |
| AB | 1622 | hCoV-19/England/ALDP-C89346/2020\|EPI_ISL_762890\|20 | ···················· | ···················· | ···················· | ···················· | ···················· | ···················· |
| AB | 1623 | hCoV-19/England/QEUH-C7FB96/2020\|EPI_ISL_762897\|20 | ···················· | ···················· | ···················· | ···················· | ···················· | ···················· |
| AB | 1624 | hCoV-19/Wales/PHWC-4A6484/2020\|EPI_ISL_765040\|2020 | ···················· | ···················· | ···················· | ···················· | ···················· | ···················· |
| AB | 1625 | hCoV-19/Wales/PHWC-4A6493/2020\|EPI_ISL_765041\|2020 | ···················· | ···················· | ···················· | ···················· | ···················· | ···················· |
| AB | 1626 | hCoV-19/Wales/PHWC-4A64A2/2020\|EPI_ISL_765042\|2020 | ···················· | ···················· | ···················· | ···················· | ···················· | ···················· |
| AB | 1627 | hCoV-19/Wales/PHWC-4A64C0/2020\|EPI_ISL_765043\|2020 | ···················· | ···················· | ···················· | ···················· | ···················· | ···················· |
| AB | 1628 | hCoV-19/Wales/PHWC-4A6509/2020\|EPI_ISL_765044\|2020 | ···················· | ···················· | ···················· | ···················· | ···················· | ···················· |
| AB | 1629 | hCoV-19/Wales/PHWC-4A6536/2020\|EPI_ISL_765045\|2020 | ···················· | ···················· | ···················· | ···················· | ···················· | ···················· |
| AB | 1631 | hCoV-19/Wales/PHWC-4A6554/2020\|EPI_ISL_765047\|2020 | ···················· | ···················· | ···················· | ···················· | ···················· | ···················· |
| AB | 1632 | hCoV-19/Wales/PHWC-4A6563/2020\|EPI_ISL_765048\|2020 | ···················· | ···················· | ···················· | ···················· | ···················· | ···················· |
| AB | 1633 | hCoV-19/Wales/PHWC-4A6572/2020\|EPI_ISL_765049\|2020 | ···················· | ···················· | ···················· | ···················· | ···················· | ···················· |
| AB | 1634 | hCoV-19/Wales/PHWC-4A6581/2020\|EPI_ISL_765050\|2020 | ···················· | ···················· | ···················· | ···················· | ···················· | ···················· |
| AB | 1635 | hCoV-19/Wales/PHWC-4A65DC/2020\|EPI_ISL_765051\|2020 | ···················· | ···················· | ···················· | ···················· | ···················· | ···················· |
| AB | 1636 | hCoV-19/Wales/PHWC-4A65FA/2020\|EPI_ISL_765052\|2020 | ···················· | ···················· | ···················· | ···················· | ···················· | ···················· |
| AB | 1637 | hCoV-19/Wales/PHWC-4A6606/2020\|EPI_ISL_765053\|2020 | ···················· | ···················· | ···················· | ···················· | ···················· | ···················· |
| AB | 1638 | hCoV-19/Wales/PHWC-4A6615/2020\|EPI_ISL_765054\|2020 | ···················· | ···················· | ···················· | ···················· | ···················· | ···················· |
| AB | 1639 | hCoV-19/Wales/PHWC-4A6624/2020\|EPI_ISL_765055\|2020 | ···················· | ···················· | ···················· | ···················· | ···················· | ···················· |
| AB | 1640 | hCoV-19/Wales/PHWC-4A6651/2020\|EPI_ISL_765056\|2020 | ···················· | ···················· | ···················· | ···················· | ···················· | ···················· |
| AB | 1641 | hCoV-19/Wales/PHWC-4A6660/2020\|EPI_ISL_765057\|2020 | ·T·················· | ···················· | ···················· | ···················· | ···················· | ···················· |
| AB | 1642 | hCoV-19/Wales/PHWC-4A667F/2020\|EPI_ISL_765058\|2020 | ···················· | ···················· | ···················· | ···················· | ···················· | ···················· |
| AB | 1643 | hCoV-19/Wales/PHWC-4A668E/2020\|EPI_ISL_765059\|2020 | ···················· | ···················· | ···················· | ···················· | ···················· | ···················· |
| AB | 1644 | hCoV-19/Wales/PHWC-4A669D/2020\|EPI_ISL_765060\|2020 | ···················· | ···················· | ···················· | ···················· | ···················· | ···················· |
| AB | 1645 | hCoV-19/Wales/PHWC-4A66AC/2020\|EPI_ISL_765061\|2020 | ···················· | ···················· | ···················· | ···················· | ···················· | ···················· |
| AB | 1646 | hCoV-19/Wales/PHWC-4A66BB/2020\|EPI_ISL_765062\|2020 | ···················· | ···················· | ···················· | ···················· | ···················· | ···················· |
| AB | 1647 | hCoV-19/Wales/PHWC-4A66CA/2020\|EPI_ISL_765063\|2020 | ···················· | ···················· | ···················· | ···················· | ···················· | ···················· |
| AB | 1649 | hCoV-19/Wales/PHWC-4A66E8/2020\|EPI_ISL_765065\|2020 | ···················· | ···················· | ···················· | ···················· | ···················· | ···················· |
| AB | 1651 | hCoV-19/Wales/PHWC-4A6703/2020\|EPI_ISL_765067\|2020 | ···················· | ···················· | ···················· | ···················· | ···················· | ···················· |
| AB | 1652 | hCoV-19/Wales/PHWC-4A6721/2020\|EPI_ISL_765068\|2020 | ···················· | ···················· | ···················· | ···················· | ···················· | ···················· |
| AB | 1653 | hCoV-19/Wales/PHWC-4A674F/2020\|EPI_ISL_765069\|2020 | ···················· | ···················· | ···················· | ···················· | ···················· | ···················· |
| AB | 1654 | hCoV-19/Wales/PHWC-4A676D/2020\|EPI_ISL_765070\|2020 | ···················· | ···················· | ···················· | ···················· | ···················· | ···················· |
| AB | 1656 | hCoV-19/Wales/PHWC-4A67A9/2020\|EPI_ISL_765072\|2020 | ···················· | ···················· | ···················· | ···················· | ···················· | ···················· |
| AB | 1657 | hCoV-19/Wales/PHWC-4A67E5/2020\|EPI_ISL_765073\|2020 | ···················· | ···················· | ···················· | ···················· | ···················· | ···················· |
| AB | 1658 | hCoV-19/Wales/PHWC-4A6800/2020\|EPI_ISL_765074\|2020 | ···················· | ···················· | ···················· | ···················· | ···················· | ···················· |
| AB | 1659 | hCoV-19/Wales/PHWC-4A682E/2020\|EPI_ISL_765075\|2020 | ···················· | ···················· | ···················· | ···················· | ·NN················· | ···················· |
| AB | 1660 | hCoV-19/Wales/PHWC-4A683D/2020\|EPI_ISL_765076\|2020 | ···················· | ···················· | ···················· | ···················· | ···················· | ···················· |
| AB | 1661 | hCoV-19/Wales/PHWC-4A684C/2020\|EPI_ISL_765077\|2020 | ···················· | ···················· | ···················· | ···················· | ···················· | ···················· |
| AB | 1662 | hCoV-19/Wales/PHWC-4A686A/2020\|EPI_ISL_765078\|2020 | ···················· | ···················· | ···················· | ···················· | ···················· | ···················· |
| AB | 1664 | hCoV-19/Wales/PHWC-4A6888/2020\|EPI_ISL_765080\|2020 | ···················· | ···················· | ···················· | ···················· | ···················· | ···················· |
| AB | 1665 | hCoV-19/Wales/PHWC-4A6897/2020\|EPI_ISL_765081\|2020 | ···················· | ···················· | ···················· | ···················· | ···················· | ···················· |
| AB | 1666 | hCoV-19/Wales/PHWC-4A68E2/2020\|EPI_ISL_765082\|2020 | ···················· | ···················· | ···················· | ···················· | ···················· | ···················· |
| AB | 1667 | hCoV-19/Wales/PHWC-4A68F1/2020\|EPI_ISL_765083\|2020 | ···················· | ···················· | ···················· | ···················· | ···················· | ···················· |
| AB | 1669 | hCoV-19/Wales/PHWC-4A692B/2020\|EPI_ISL_765085\|2020 | ···················· | ···················· | ···················· | ···················· | ···················· | ···················· |
| AB | 1670 | hCoV-19/Wales/PHWC-4A693A/2020\|EPI_ISL_765086\|2020 | ···················· | ···················· | ···················· | ···················· | ···················· | ···················· |
| AB | 1671 | hCoV-19/Wales/PHWC-4A6967/2020\|EPI_ISL_765087\|2020 | ···················· | ···················· | ···················· | ···················· | ···················· | ···················· |
| AB | 1673 | hCoV-19/Wales/PHWC-4A6994/2020\|EPI_ISL_765089\|2020 | ···················· | ···················· | ···················· | ···················· | ···················· | ···················· |
| AB | 1674 | hCoV-19/Wales/PHWC-4A69A3/2020\|EPI_ISL_765090\|2020 | ···················· | ···················· | ···················· | ···················· | ···················· | ···················· |
| AB | 1675 | hCoV-19/Wales/PHWC-4A69B2/2020\|EPI_ISL_765091\|2020 | ···················· | ···················· | ···················· | ···················· | ···················· | ···················· |
| AB | 1678 | hCoV-19/Wales/PHWC-4A6A0A/2020\|EPI_ISL_765094\|2020 | ···················· | ···················· | ···················· | ···················· | ···················· | ···················· |
| AB | 1679 | hCoV-19/Wales/PHWC-4A6A37/2020\|EPI_ISL_765095\|2020 | ···················· | ···················· | ···················· | ···················· | ···················· | ···················· |
| AB | 1680 | hCoV-19/Wales/PHWC-4A6A46/2020\|EPI_ISL_765096\|2020 | ···················· | ···················· | ···················· | ···················· | ···················· | ···················· |
| AB | 1681 | hCoV-19/Wales/PHWC-4A6A64/2020\|EPI_ISL_765097\|2020 | ···················· | ···················· | ···················· | ···················· | ···················· | ···················· |
| AB | 1682 | hCoV-19/Wales/PHWC-4A6A73/2020\|EPI_ISL_765098\|2020 | ···················· | ···················· | ···················· | ···················· | ···················· | ···················· |
| AB | 1683 | hCoV-19/Wales/PHWC-4A6AA0/2020\|EPI_ISL_765099\|2020 | ···················· | ···················· | ···················· | ···················· | ···················· | ···················· |
| AB | 1684 | hCoV-19/England/205121137/2020\|EPI_ISL_766317\|2020 | ···················· | ···················· | ···················· | ···················· | ···················· | ···················· |
| AB | 1685 | hCoV-19/Gibraltar/205121188/2020\|EPI_ISL_766318\|20 | ···················· | ···················· | ···················· | ···················· | ···················· | ···················· |
| AB | 1686 | hCoV-19/Gibraltar/205121189/2020\|EPI_ISL_766319\|20 | ···················· | ···················· | ···················· | ···················· | ···················· | ···················· |
| AB | 1687 | hCoV-19/Gibraltar/205121248/2020\|EPI_ISL_766370\|20 | ···················· | ···················· | ···················· | ···················· | ···················· | ···················· |
| AB | 1688 | hCoV-19/Gibraltar/205121251/2020\|EPI_ISL_766371\|20 | ···················· | ···················· | ···················· | ···················· | ···················· | ···················· |
| AB | 1689 | hCoV-19/Gibraltar/205121252/2020\|EPI_ISL_766372\|20 | ···················· | ···················· | ···················· | ···················· | ···················· | ···················· |
| AB | 1690 | hCoV-19/Gibraltar/205121253/2020\|EPI_ISL_766373\|20 | ···················· | ···················· | ···················· | ···················· | ···················· | ···················· |
| AB | 1691 | hCoV-19/Gibraltar/205121255/2020\|EPI_ISL_766374\|20 | ···················· | ···················· | ···················· | ···················· | ···················· | ···················· |
| AB | 1692 | hCoV-19/Gibraltar/205121256/2020\|EPI_ISL_766375\|20 | ···················· | ·A·················· | ···················· | ···················· | ···················· | ···················· |
| AB | 1693 | hCoV-19/Gibraltar/205121257/2020\|EPI_ISL_766376\|20 | ···················· | ···················· | ···················· | ···················· | ···················· | ···················· |
| AB | 1694 | hCoV-19/Gibraltar/205121258/2020\|EPI_ISL_766377\|20 | ···················· | ···················· | ···················· | ···················· | ···················· | ···················· |
| AB | 1695 | hCoV-19/Gibraltar/205121259/2020\|EPI_ISL_766378\|20 | ···················· | ···················· | ···················· | ···················· | ···················· | ···················· |
| AB | 1696 | hCoV-19/Gibraltar/205121260/2020\|EPI_ISL_766379\|20 | ···················· | ···················· | ···················· | ···················· | ···················· | ···················· |
| AB | 1697 | hCoV-19/Gibraltar/205121261/2020\|EPI_ISL_766380\|20 | ···················· | ···················· | ···················· | ···················· | ···················· | ···············A···· |

| | | | | | | | |
|---|---|---|---|---|---|---|---|
| AB | 1698 hCoV-19/Gibraltar/205121262/2020\|EPI_ISL_766381\|20 | ···················· | ···················· | ···················· | ···················· | ···················· | ···················· |
| AB | 1699 hCoV-19/Gibraltar/205121263/2020\|EPI_ISL_766382\|20 | ···················· | ···················· | ···················· | ···················· | ···················· | ···················· |
| AB | 1700 hCoV-19/Gibraltar/205121264/2020\|EPI_ISL_766383\|20 | ···················· | ···················· | ···················· | ···················· | ···················· | ···················· |
| AB | 1701 hCoV-19/Gibraltar/205121265/2020\|EPI_ISL_766384\|20 | ···················· | ···················· | ···················· | ···················· | ···················· | ···················· |
| AB | 1702 hCoV-19/Gibraltar/205121266/2020\|EPI_ISL_766385\|20 | ···················· | ···················· | ···················· | ···················· | ···················· | ···················· |
| AB | 1703 hCoV-19/Gibraltar/205121267/2020\|EPI_ISL_766386\|20 | ···················· | ···················· | ···················· | ···················· | ···················· | ···················· |
| AB | 1704 hCoV-19/Gibraltar/205121268/2020\|EPI_ISL_766387\|20 | ···················· | ···················· | ···················· | ···················· | ···················· | ···················· |
| AB | 1705 hCoV-19/Gibraltar/205121269/2020\|EPI_ISL_766388\|20 | ···················· | ···················· | ···················· | ···················· | ···················· | ···················· |
| AB | 1706 hCoV-19/Gibraltar/205121270/2020\|EPI_ISL_766389\|20 | ···················· | ···················· | ···················· | ···················· | ···················· | ···················· |
| AB | 1707 hCoV-19/Gibraltar/205121271/2020\|EPI_ISL_766390\|20 | ···················· | ···················· | ···················· | ···················· | ···················· | ···················· |
| AB | 1708 hCoV-19/Gibraltar/205121272/2020\|EPI_ISL_766391\|20 | ···················· | ···················· | ···················· | ···················· | ···················· | ···············A···· |
| AB | 1709 hCoV-19/Gibraltar/205121273/2020\|EPI_ISL_766392\|20 | ···················· | ···················· | ···················· | ···················· | ···················· | ···················· |
| AB | 1710 hCoV-19/Gibraltar/205121275/2020\|EPI_ISL_766393\|20 | ···················· | ···················· | ···················· | ···················· | ···················· | ···············A···· |
| AB | 1711 hCoV-19/Gibraltar/205121276/2020\|EPI_ISL_766394\|20 | ···················· | ···················· | ···················· | ···················· | ···················· | ···················· |
| AB | 1712 hCoV-19/England/205121330/2020\|EPI_ISL_766395\|2020 | ···················· | ···················· | ···················· | ···················· | ···················· | ···················· |
| AB | 1713 hCoV-19/England/205140691/2020\|EPI_ISL_766396\|2020 | ···················· | ···················· | ···················· | ···················· | ···················· | ···················· |
| AB | 1714 hCoV-19/England/205140692/2020\|EPI_ISL_766397\|2020 | ···················· | ·A·················· | ···················· | ···················· | ···················· | ···················· |
| AB | 1715 hCoV-19/England/205140694/2020\|EPI_ISL_766398\|2020 | ···················· | ···················· | ···················· | ···················· | ···················· | ···················· |
| AB | 1716 hCoV-19/England/205140698/2020\|EPI_ISL_766399\|2020 | ···················· | ···················· | ···················· | ···················· | ······T············· | ···················· |
| AB | 1717 hCoV-19/Bangladesh/DNAS_DRICM_CPH_isl_441_BGD/2020 | ···················· | ···················· | ···················· | ···················· | ···················· | ···················· |
| AB | 1718 hCoV-19/Bangladesh/DNAS_DRCIM_CPH_isl_475_BGD/2020 | ···················· | ···················· | ···················· | ···················· | ···················· | ···················· |
| AB | 1719 hCoV-19/Bangladesh/DNAS_DRCIM_CPH_isl_428_BGD/2020 | ···················· | ···················· | ···················· | ···················· | ···················· | ···················· |
| AB | 1720 hCoV-19/Bangladesh/DNAS_DRICM_CPH_isl_521_BGD/2020 | ···················· | ···················· | ···················· | ···················· | ···················· | ···················· |
| AB | 1721 hCoV-19/Bangladesh/DNAS_DRCIM_P_isl_T_BGD/2020\|EPI | ···················· | ···················· | ···················· | ···················· | ···················· | ···················· |
| AB | 1722 hCoV-19/Bangladesh/DNAS_DRCIM_CPH_isl_417_BGD/2020 | ···················· | ···················· | ···················· | ···················· | ···················· | ···················· |
| AB | 1723 hCoV-19/Bangladesh/DNAS_DRCIM_CPH_isl_485_BGD/2020 | ···················· | ···················· | ···················· | ···················· | ···················· | ···················· |
| AB | 1724 hCoV-19/Bangladesh/DNAS_DRCIM_CPH_isl_509_BGD/2020 | ···················· | ···················· | ···················· | ···················· | ···················· | ···················· |
| AB | 1725 hCoV-19/Bangladesh/DNAS_DRCIM_CPH_isl_479_BGD/2020 | ···················· | ···················· | ···················· | ···················· | ···················· | ···················· |
| AB | 1726 hCoV-19/Bangladesh/DNAS_DRCIM_CPH_isl_486_BGD/2020 | ···················· | ···················· | ···················· | ···················· | ···················· | ···················· |
| AB | 1727 hCoV-19/Bangladesh/DRICM_DNAS_CPH_isl_430_BGD/2020 | ···················· | ···················· | ···················· | ···················· | ···················· | ···················· |
| AB | 1728 hCoV-19/Bangladesh/DNAS_DRCIM_CPH_isl_510_BGD/2020 | ···················· | ···················· | ···················· | ···················· | ···················· | ···················· |
| AB | 1729 hCoV-19/Bangladesh/DNAS_DRCIM_CPH_isl_506_BGD/2020 | ···················· | ···················· | ···················· | ···················· | ···················· | ···················· |
| AB | 1730 hCoV-19/Bangladesh/DNAS_DRCIM_CPH_isl_438_BGD/2020 | ···················· | ···················· | ···················· | ···················· | ···················· | ···················· |
| AB | 1731 hCoV-19/Bangladesh/DNAS_DRCIM_CPH_isl_525_BGD/2020 | ···················· | ···················· | ···················· | ···················· | ···················· | ···················· |
| AB | 1732 hCoV-19/Bangladesh/DNAS_DRCIM_CPH_isl_515_BGD/2020 | ···················· | ···················· | ···················· | ···················· | ···················· | ···················· |
| AB | 1733 hCoV-19/Bangladesh/DNAS_DRCIM_BSH_isl_1_BGD/2020\|E | ···················· | ···················· | ···················· | ···················· | ···················· | ···················· |
| AB | 1734 hCoV-19/Bangladesh/DNAS_DRCIM_RAB_isl_88_BGD/2020\| | ···················· | ···················· | ···················· | ···················· | ···················· | ···················· |
| AB | 1735 hCoV-19/Bangladesh/DNAS_DRCIM_RAB_isl_86_BGD/2020\| | ···················· | ···················· | ···················· | ···················· | ···················· | ···················· |
| AB | 1736 hCoV-19/Bangladesh/DNAS_DRCIM_CPH_isl_426_BGD/2020 | ···················· | ···················· | ···················· | ···················· | ···················· | ···················· |
| AB | 1737 hCoV-19/Bangladesh/DNAS_DRCIM_CPH_isl_461_BGD/2020 | ···················· | ···················· | ···················· | ···················· | ···················· | ···················· |
| AB | 1738 hCoV-19/Bangladesh/DNAS_DRCIM_CPH_isl_437_BGD/2020 | ···················· | ···················· | ···················· | ···················· | ···················· | ···················· |
| AB | 1739 hCoV-19/Bangladesh/DNAS_DRCIM_RAB_isl_114_BGD/2020 | ···················· | ···················· | ···················· | ···················· | ···················· | ···················· |
| AB | 1740 hCoV-19/Bangladesh/DNAS_DRCIM_RAB_isl_118_BGD/2020 | ···················· | ···················· | ···················· | ···················· | ···················· | ···················· |
| AB | 1741 hCoV-19/Bangladesh/DNAS_DRCIM_CPH_isl_537_BGD/2020 | ···················· | ···················· | ···················· | ···················· | ···················· | ···················· |
| AB | 1742 hCoV-19/Bangladesh/DNAS_DRCIM_BSH_isl_9_BGD/2020\|E | ···················· | ···················· | ···················· | ···················· | ···················· | ···················· |
| AB | 1743 hCoV-19/Bangladesh/DNAS_DRCIM_isl_113/2020\|EPI_ISL | ···················· | ···················· | ···················· | ···················· | ···················· | ···················· |
| AB | 1744 hCoV-19/England/ALDP-C8BA5E/2020\|EPI_ISL_777020\|20 | ···················· | ···················· | ···················· | ···················· | ···················· | ···················· |
| AB | 1745 hCoV-19/England/QEUH-C93A62/2020\|EPI_ISL_777021\|20 | ···················· | ···················· | ···················· | ···················· | ···················· | ···················· |
| AB | 1746 hCoV-19/England/ALDP-C8BAB8/2020\|EPI_ISL_777024\|20 | ···················· | ···················· | ···················· | ···················· | ···················· | ···················· |
| AB | 1747 hCoV-19/England/ALDP-C8C11C/2020\|EPI_ISL_777027\|20 | ···················· | ·A·················· | ···················· | ···················· | ···················· | ···A················ |
| AB | 1748 hCoV-19/England/ALDP-C8F2E9/2020\|EPI_ISL_777033\|20 | ···················· | ···················· | ···················· | ···················· | ···················· | ···················· |
| AB | 1749 hCoV-19/England/ALDP-C8C1A3/2020\|EPI_ISL_777036\|20 | ···················· | ···················· | ···················· | ···················· | ···················· | ···················· |
| AB | 1750 hCoV-19/England/QEUH-C93868/2020\|EPI_ISL_777038\|20 | ···················· | ···················· | ···················· | ···················· | ···················· | ···················· |
| AB | 1751 hCoV-19/England/MILK-C8ECCE/2020\|EPI_ISL_777039\|20 | ···················· | ···················· | ···················· | ···················· | ···················· | ···A················ |
| AB | 1752 hCoV-19/England/ALDP-C8F35F/2020\|EPI_ISL_777042\|20 | ···················· | ·A·················· | ···················· | ···················· | ···················· | ···················· |
| AB | 1753 hCoV-19/England/ALDP-C8D0E1/2020\|EPI_ISL_777043\|20 | ···················· | ···················· | ···················· | ···················· | ···················· | ···················· |
| AB | 1754 hCoV-19/England/ALDP-C8CCEE/2020\|EPI_ISL_777044\|20 | ···················· | ···················· | ···················· | ···················· | ···················· | ···················· |
| AB | 1755 hCoV-19/England/ALDP-C8D6D0/2020\|EPI_ISL_777046\|20 | ···················· | ···················· | ···················· | ···················· | ···················· | ···················· |
| AB | 1756 hCoV-19/England/QEUH-C93A53/2020\|EPI_ISL_777047\|20 | ···················· | ···················· | ···················· | ···················· | ···················· | ···················· |
| AB | 1757 hCoV-19/England/ALDP-C8C3F8/2020\|EPI_ISL_777048\|20 | ···················· | ···················· | ···················· | ···················· | ···················· | ···················· |
| AB | 1758 hCoV-19/England/ALDP-C8D157/2020\|EPI_ISL_777049\|20 | ···················· | ···················· | ···················· | ···················· | ···················· | ···················· |
| AB | 1759 hCoV-19/England/ALDP-C8C3E9/2020\|EPI_ISL_777051\|20 | ···················· | ···················· | ···················· | ···················· | ···················· | ···················· |
| AB | 1760 hCoV-19/England/ALDP-C8BAC7/2020\|EPI_ISL_777052\|20 | ···················· | ···················· | ···················· | ···················· | ···················· | ···················· |
| AB | 1761 hCoV-19/England/ALDP-C8CCDF/2020\|EPI_ISL_777053\|20 | ···················· | ···················· | ···················· | ···················· | ···················· | ···················· |
| AB | 1762 hCoV-19/England/MILK-C90458/2020\|EPI_ISL_777056\|20 | ···················· | ·A·················· | ···················· | ···················· | ···················· | ···A················ |
| AB | 1763 hCoV-19/England/QEUH-C938E0/2020\|EPI_ISL_777059\|20 | ···················· | ···················· | ···················· | ···················· | ···················· | ···················· |
| AB | 1764 hCoV-19/England/ALDP-C8C589/2020\|EPI_ISL_777060\|20 | ···················· | ···················· | ···················· | ···················· | ···················· | ···················· |
| AB | 1765 hCoV-19/England/ALDP-C9050A/2020\|EPI_ISL_777063\|20 | ···················· | ···················· | ···················· | ···················· | ···················· | ···················· |
| AB | 1766 hCoV-19/England/ALDP-C8F2CB/2020\|EPI_ISL_777067\|20 | ·····G·············· | ···················· | ···················· | ···················· | ···················· | ···················· |
| AB | 1767 hCoV-19/England/ALDP-C8C8F9/2020\|EPI_ISL_777071\|20 | ···················· | ···················· | ···················· | ···················· | ···················· | ···················· |
| AB | 1768 hCoV-19/England/ALDP-C8BAF4/2020\|EPI_ISL_777072\|20 | ···················· | ···················· | ···················· | ···················· | ···················· | ···················· |
| AB | 1769 hCoV-19/England/ALDP-C8C97E/2020\|EPI_ISL_777077\|20 | ···················· | ···················· | ···················· | ···················· | ···················· | ···················· |
| AB | 1770 hCoV-19/England/ALDP-C8D685/2020\|EPI_ISL_777078\|20 | ···················· | ···················· | ···················· | ···················· | ···················· | ···················· |
| AB | 1771 hCoV-19/England/ALDP-C8CCA2/2020\|EPI_ISL_777079\|20 | ···················· | ···················· | ···················· | ···················· | ···················· | ···················· |
| AB | 1772 hCoV-19/England/MILK-C90968/2020\|EPI_ISL_777080\|20 | ···················· | ···················· | ···················· | ···················· | ···················· | ···················· |
| AB | 1773 hCoV-19/England/QEUH-C901AC/2020\|EPI_ISL_777081\|20 | ···················· | ···················· | ···················· | ···················· | ···················· | ···················· |
| AB | 1774 hCoV-19/England/ALDP-C8FDD9/2020\|EPI_ISL_777083\|20 | ···················· | ···················· | ···················· | ···················· | ···················· | ···················· |
| AB | 1775 hCoV-19/England/MILK-C8EF98/2020\|EPI_ISL_777085\|20 | ···················· | ···················· | ···················· | ···················· | ···················· | ···················· |
| AB | 1776 hCoV-19/England/QEUH-C936C8/2020\|EPI_ISL_777089\|20 | ···················· | ···················· | ···················· | ···················· | ···················· | ···················· |
| AB | 1777 hCoV-19/England/ALDP-C8CD72/2020\|EPI_ISL_777091\|20 | ···················· | ···················· | ···················· | ···················· | ···················· | ···················· |

```
AB    1778 hCoV-19/England/MILK-C6C622/2020|EPI_ISL_778350|20    ····················  ····················  ·····················  ····················  ····················  ····················
AB    1779 hCoV-19/England/QEUH-C65489/2020|EPI_ISL_778356|20    ····················  ····················  ·····················  ····················  ····················  ····················
AB    1780 hCoV-19/England/ALDP-C6B77B/2020|EPI_ISL_778357|20    ····················  ····················  ·····················  ····················  ····················  ····················
AB    1781 hCoV-19/England/MILK-C6F70E/2020|EPI_ISL_778369|20    ····················  ····················  ·····················  ····················  ····················  ····················
AB    1782 hCoV-19/England/MILK-C6C9CF/2020|EPI_ISL_778370|20    ····················  ····················  ·····················  ····················  ····················  ····················
AB    1783 hCoV-19/England/MILK-C6CF27/2020|EPI_ISL_778371|20    ····················  ·A··················  ·····················  ····················  ····················  ···A················
AB    1784 hCoV-19/England/QEUH-C643E7/2020|EPI_ISL_778372|20    ····················  ····················  ·····················  ····················  ····················  ····················
AB    1785 hCoV-19/England/QEUH-C65410/2020|EPI_ISL_778377|20    ····················  ····················  ·····················  ····················  ····················  ····················
AB    1786 hCoV-19/England/MILK-C6CC8A/2020|EPI_ISL_778383|20    ····················  ····················  ·····················  ····················  ····················  ····················
AB    1787 hCoV-19/England/MILK-C6F56E/2020|EPI_ISL_778386|20    ····················  ····················  ···········T·········  ····················  ····················  ····················
AB    1788 hCoV-19/England/ALDP-C6A141/2020|EPI_ISL_778390|20    ····················  ····················  ·····················  ····················  ····················  ····················
AB    1789 hCoV-19/England/ALDP-C6B7D5/2020|EPI_ISL_778392|20    ····················  ····················  ·····················  ····················  ····················  ····················
AB    1790 hCoV-19/England/MILK-C6F65C/2020|EPI_ISL_778393|20    ····················  ····················  ·····················  ····················  ····················  ····················
AB    1791 hCoV-19/England/ALDP-C6B8F0/2020|EPI_ISL_778396|20    ····················  ····················  ·····················  ····················  ····················  ····················
AB    1792 hCoV-19/England/MILK-C6F66B/2020|EPI_ISL_778398|20    ····················  ····················  ·····················  ····················  ····················  ····················
AB    1793 hCoV-19/South Korea/KDCA0429/2020|EPI_ISL_760213|2    ····················  ····················  ·····················  ····················  ····················  ····················
AB    1794 hCoV-19/South Korea/KDCA0430/2020|EPI_ISL_760214|2    ····················  ····················  ·····················  ····················  ····················  ····················
AB    1795 hCoV-19/South Korea/KDCA0431/2020|EPI_ISL_760215|2    ····················  ····················  ·····················  ····················  ····················  ····················
AB    1796 hCoV-19/England/QEUH-C93DA5/2020|EPI_ISL_762011|20    ····················  ····················  ·····················  ····················  ······T·············  ····················
AB    1797 hCoV-19/England/CAMC-C997ED/2020|EPI_ISL_762015|20    ····················  ····················  ·····················  ····················  ····················  ····················
AB    1798 hCoV-19/England/QEUH-C935CB/2020|EPI_ISL_762017|20    ····················  ·A··················  ·····················  ····················  ····················  ···A················
AB    1799 hCoV-19/England/QEUH-C93042/2020|EPI_ISL_762021|20    ····················  ····················  ·····················  ····················  ······T·············  ····················
AB    1800 hCoV-19/England/QEUH-C9321F/2020|EPI_ISL_762022|20    ····················  ····················  ·····················  ····················  ····················  ····················
AB    1801 hCoV-19/England/CAMC-C9902E/2020|EPI_ISL_762024|20    ····················  ····················  ·····················  ····················  ····················  ····················
AB    1802 hCoV-19/England/CAMC-C98F6E/2020|EPI_ISL_762025|20    ····················  ····················  ·····················  ····················  ····················  ····················
AB    1803 hCoV-19/England/CAMC-C9961D/2020|EPI_ISL_762026|20    ····················  ····················  ·····················  ····················  ····················  ····················
AB    1804 hCoV-19/England/QEUH-C934BF/2020|EPI_ISL_762039|20    ····················  ····················  ·····················  ····················  ····················  ····················
AB    1805 hCoV-19/England/CAMC-C99501/2020|EPI_ISL_762041|20    ····················  ····················  ·····················  ····················  ····················  ····················
AB    1806 hCoV-19/England/CAMC-C994F5/2020|EPI_ISL_762042|20    ····················  ····················  ·····················  ····················  ····················  ····················
AB    1807 hCoV-19/England/QEUH-C92D2E/2020|EPI_ISL_762047|20    ····················  ····················  ·····················  ····················  ····················  ····················
AB    1808 hCoV-19/England/QEUH-C7C74A/2020|EPI_ISL_762877|20    ····················  ····················  ·····················  ····················  ····················  ····················
AB    1809 hCoV-19/Netherlands/ZH-EMC-1231/2020|EPI_ISL_76330    ····················  ····················  ·····················  ····················  ····················  ····················
AB    1810 hCoV-19/Netherlands/ZH-EMC-1232/2020|EPI_ISL_76330    ····················  ······A·············  ·····················  ····················  ····················  ····················
AB    1811 hCoV-19/Netherlands/ZH-EMC-1234/2020|EPI_ISL_76330    ····················  ····················  ·····················  ····················  ····················  ····················
AB    1812 hCoV-19/Netherlands/ZH-EMC-1236/2020|EPI_ISL_76330    ····················  ····················  ·····················  ····················  ····················  ····················
AB    1813 hCoV-19/Netherlands/ZH-EMC-1237/2020|EPI_ISL_76330    ····················  ····················  ·····················  ····················  ····················  ····················
AB    1814 hCoV-19/Netherlands/ZH-EMC-1238/2020|EPI_ISL_76330    ····················  ····················  ·····················  ····················  ····················  ····················
AB    1815 hCoV-19/Netherlands/ZH-EMC-1239/2020|EPI_ISL_76330    ····················  ····················  ·····················  ····················  ····················  ····················
AB    1816 hCoV-19/Netherlands/ZH-EMC-1240/2020|EPI_ISL_76330    ····················  ····················  ·····················  ····················  ····················  ····················
AB    1817 hCoV-19/Netherlands/ZH-EMC-1241/2020|EPI_ISL_76331    ····················  G···················  ·····················  ····················  ····················  ····················
AB    1818 hCoV-19/Netherlands/ZH-EMC-1242/2020|EPI_ISL_76331    ····················  ····················  ·····················  ····················  ····················  ····················
AB    1819 hCoV-19/Netherlands/ZH-EMC-1244/2020|EPI_ISL_76331    ····················  ····················  ·····················  ····················  ······T·············  ····················
AB    1820 hCoV-19/Netherlands/ZH-EMC-1245/2020|EPI_ISL_76331    ····················  ····················  ·····················  ····················  ····················  ····················
AB    1821 hCoV-19/Netherlands/ZH-EMC-1246/2020|EPI_ISL_76331    ····················  ····················  ·····················  ····················  ····················  ····················
AB    1822 hCoV-19/Italy/UMB-IZSUM-48682-25/2020|EPI_ISL_7633    ····················  ····················  ·····················  ····················  ····················  ····················
AB    1823 hCoV-19/USA/OH-ODH-SC35004/2020|EPI_ISL_763319|202    ····················  ····················  ·····················  ····················  ····················  ····················
AB    1824 hCoV-19/Italy/UMB-IZSUM-48682-19/2020|EPI_ISL_7633    ····················  ····················  ·····················  ····················  ····················  ····················
AB    1825 hCoV-19/USA/OH-ODH-SC35005/2020|EPI_ISL_763321|202    ····················  ····················  ·····················  ····················  ····················  ····················
AB    1826 hCoV-19/Italy/UMB-IZSUM-48682-14/2020|EPI_ISL_7633    ····················  ····················  ·····················  ····················  ····················  ····················
AB    1827 hCoV-19/Italy/UMB-IZSUM-48682-13/2020|EPI_ISL_7633    ····················  ····················  ·····················  ····················  ····················  ····················
AB    1828 hCoV-19/Italy/UMB-IZSUM-48682-6/2020|EPI_ISL_76332    ····················  ····················  ·····················  ····················  ····················  ····················
AB    1829 hCoV-19/USA/FL-BPHL-2259/2020|EPI_ISL_763342|2020-    ····················  ····················  ·····················  ····················  ····················  ····················
AB    1830 hCoV-19/Netherlands/ZH-EMC-1219/2020|EPI_ISL_76334    ····················  ····················  ·····················  ····················  ····················  ····················
AB    1831 hCoV-19/Netherlands/ZH-EMC-1130/2020|EPI_ISL_76334    ····················  ····················  ·····················  ····················  ····················  ····················
AB    1832 hCoV-19/Ireland/LD-NVRL-20G48731/2020|EPI_ISL_7633    ····················  ····················  ·····················  ····················  ····················  ····················
AB    1833 hCoV-19/Ireland/WH-NVRL-20Z04384/2020|EPI_ISL_7633    ····················  ····················  ·····················  ····················  ····················  ····················
AB    1834 hCoV-19/Netherlands/ZH-EMC-1192/2020|EPI_ISL_76335    ····················  ····················  ·····················  ····················  ····················  ····················
AB    1835 hCoV-19/Netherlands/ZH-EMC-1207/2020|EPI_ISL_76335    ····················  ····················  ·····················  ····················  ····················  ····················
AB    1836 hCoV-19/Netherlands/ZH-EMC-1210/2020|EPI_ISL_76335    ····················  ····················  ·····················  ····················  ····················  ····················
AB    1837 hCoV-19/Netherlands/ZH-EMC-1181/2020|EPI_ISL_76335    ····················  ····················  ·····················  ····················  ····················  ····················
AB    1838 hCoV-19/Netherlands/OV-EMC-127/2020|EPI_ISL_763359    ····················  ····················  ·····················  ····················  ····················  ····················
AB    1839 hCoV-19/Wales/PHWC-4A3D31/2020|EPI_ISL_764623|2020    ····················  ····················  ·····················  ····················  ····················  ····················
AB    1841 hCoV-19/Wales/PHWC-4A3D5F/2020|EPI_ISL_764625|2020    ····················  ····················  ·····················  ····················  ····················  ····················
AB    1842 hCoV-19/Wales/PHWC-4A3D6E/2020|EPI_ISL_764626|2020    ····················  ····················  ·····················  ····················  ····················  ····················
AB    1843 hCoV-19/Wales/PHWC-4A3D7D/2020|EPI_ISL_764627|2020    ····················  ····················  ·····················  ····················  ····················  ····················
AB    1844 hCoV-19/Wales/PHWC-4A3D8C/2020|EPI_ISL_764628|2020    ····················  ····················  ·····················  ····················  ····················  ····················
AB    1845 hCoV-19/Wales/PHWC-4A3D9B/2020|EPI_ISL_764629|2020    ····················  ····················  ·····················  ····················  ····················  ····················
AB    1846 hCoV-19/Wales/PHWC-4A3DD7/2020|EPI_ISL_764630|2020    ····················  ····················  ·····················  ····················  ····················  ····················
AB    1847 hCoV-19/Wales/PHWC-4A3DE6/2020|EPI_ISL_764631|2020    ····················  ····················  ·····················  ····················  ····················  ····················
AB    1848 hCoV-19/Wales/PHWC-4A3E2F/2020|EPI_ISL_764632|2020    ····················  ····················  ·····················  ····················  ····················  ····················
AB    1849 hCoV-19/Wales/PHWC-4A3E3E/2020|EPI_ISL_764633|2020    ····················  ····················  ·····················  ····················  ····················  ····················
AB    1850 hCoV-19/Wales/PHWC-4A3E4D/2020|EPI_ISL_764634|2020    ····················  ····················  ·····················  ····················  ····················  ····················
AB    1851 hCoV-19/Wales/PHWC-4A3E5C/2020|EPI_ISL_764635|2020    ····················  ····················  ·····················  ····················  ····················  ····················
AB    1853 hCoV-19/Wales/PHWC-4A3E89/2020|EPI_ISL_764637|2020    ····················  ····················  ·····················  ····················  ····················  ····················
AB    1854 hCoV-19/Wales/PHWC-4A3E98/2020|EPI_ISL_764638|2020    ····················  ····················  ·····················  ····················  ····················  ····················
AB    1855 hCoV-19/Wales/PHWC-4A3EA7/2020|EPI_ISL_764639|2020    ····················  ····················  ·····················  ····················  ····················  ····················
AB    1856 hCoV-19/Wales/PHWC-4A3EB6/2020|EPI_ISL_764640|2020    ····················  ····················  ·····················  ····················  ····················  ····················
AB    1857 hCoV-19/Wales/PHWC-4A3EE3/2020|EPI_ISL_764641|2020    ····················  ····················  ·····················  ····················  ····················  ····················
AB    1858 hCoV-19/Wales/PHWC-4A3F0E/2020|EPI_ISL_764642|2020    ····················  ····················  ·····················  ····················  ····················  ····················
AB    1859 hCoV-19/Wales/PHWC-4A3F1D/2020|EPI_ISL_764643|2020    ····················  ····················  ·····················  ····················  ····················  ····················
```

| | | | | | | | | |
|---|---|---|---|---|---|---|---|---|
| AB | 1860 | hCoV-19/Wales/PHWC-4A3F2C/2020\|EPI_ISL_764644\|2020 | ···················· | ···················· | ···················· | ···················· | ···················· | ···················· |
| AB | 1861 | hCoV-19/Wales/PHWC-4A3F3B/2020\|EPI_ISL_764645\|2020 | ···················· | ···················· | ···················· | ···················· | ···················· | ···················· |
| AB | 1862 | hCoV-19/Wales/PHWC-4A3F59/2020\|EPI_ISL_764646\|2020 | ···················· | ···················· | ···················· | ···················· | ···················· | ···················· |
| AB | 1863 | hCoV-19/Wales/PHWC-4A3F77/2020\|EPI_ISL_764647\|2020 | ···················· | ···················· | ···················· | ···················· | ···················· | ···················· |
| AB | 1866 | hCoV-19/Wales/PHWC-4A3FC2/2020\|EPI_ISL_764650\|2020 | ···················· | ···················· | ···················· | ···················· | ···················· | ···················· |
| AB | 1868 | hCoV-19/Wales/PHWC-4A4019/2020\|EPI_ISL_764652\|2020 | ···················· | ···················· | ···················· | ···················· | ···················· | ···················· |
| AB | 1873 | hCoV-19/Wales/PHWC-4A4222/2020\|EPI_ISL_764668\|2020 | ···················· | ···················· | ···················· | ···················· | ···················· | ···················· |
| AB | 1877 | hCoV-19/Wales/PHWC-4A427D/2020\|EPI_ISL_764672\|2020 | ···················· | ···················· | ···················· | ···················· | ···················· | ···················· |
| AB | 1882 | hCoV-19/Wales/PHWC-4A42E6/2020\|EPI_ISL_764677\|2020 | ···················· | ···················· | ···················· | ···················· | ···················· | ···················· |
| AB | 1883 | hCoV-19/Wales/PHWC-4A42F5/2020\|EPI_ISL_764678\|2020 | ···················· | ···················· | ···················· | ···················· | ···················· | ···················· |
| AB | 1884 | hCoV-19/Wales/PHWC-4A4301/2020\|EPI_ISL_764679\|2020 | ···················· | ···················· | ···················· | ···················· | ···················· | ···················· |
| AB | 1887 | hCoV-19/Wales/PHWC-4A435C/2020\|EPI_ISL_764682\|2020 | ···················· | ···················· | ···················· | ···················· | ···················· | ···················· |
| AB | 1888 | hCoV-19/Wales/PHWC-4A437A/2020\|EPI_ISL_764683\|2020 | ···················· | ···················· | ···················· | ···················· | ···················· | ···················· |
| AB | 1889 | hCoV-19/Wales/PHWC-4A4389/2020\|EPI_ISL_764684\|2020 | ···················· | ···················· | ···················· | ···················· | ···················· | ···················· |
| AB | 1890 | hCoV-19/Wales/PHWC-4A4398/2020\|EPI_ISL_764685\|2020 | ···················· | ···················· | ···················· | ···················· | ···················· | ···················· |
| AB | 1891 | hCoV-19/Wales/PHWC-4A43B6/2020\|EPI_ISL_764686\|2020 | ···················· | ···················· | ···················· | ···················· | ···················· | ···················· |
| AB | 1893 | hCoV-19/Wales/PHWC-4A43D4/2020\|EPI_ISL_764688\|2020 | ···················· | ···················· | ···················· | ···················· | ···················· | ···················· |
| AB | 1895 | hCoV-19/Wales/PHWC-4A43F2/2020\|EPI_ISL_764690\|2020 | ···················· | ···················· | ···················· | ···················· | ···················· | ···················· |
| AB | 1899 | hCoV-19/Wales/PHWC-4A4459/2020\|EPI_ISL_764694\|2020 | ···················· | ···················· | ···················· | ···················· | ···················· | ···················· |
| AB | 1904 | hCoV-19/Wales/PHWC-4A44B3/2020\|EPI_ISL_764699\|2020 | ···················· | ···················· | ···················· | ···················· | ···················· | ···················· |
| AB | 1905 | hCoV-19/USA/AZ-WL-CVG00023/2020\|EPI_ISL_765955\|202 | ···················· | ·A·················· | ···················· | ···················· | ···················· | ···················· |
| AB | 1906 | hCoV-19/USA/WY-WPHL-20161403/2020\|EPI_ISL_765960\|2 | ···················· | ···················· | ···················· | ···················· | ···················· | ···················· |
| AB | 1907 | hCoV-19/USA/WY-WPHL-20160562/2020\|EPI_ISL_765961\|2 | ···················· | ···················· | ···················· | ···················· | ···················· | ···················· |
| AB | 1908 | hCoV-19/USA/WY-WPHL-20160575/2020\|EPI_ISL_765962\|2 | ···················· | ···················· | ···················· | ···················· | ···················· | ···················· |
| AB | 1909 | hCoV-19/USA/WY-WPHL-20160752/2020\|EPI_ISL_765963\|2 | ···················· | ·A·················· | ···················· | ···················· | ···················· | ···················· |
| AB | 1910 | hCoV-19/USA/WY-WPHL-20160842/2020\|EPI_ISL_765964\|2 | ···················· | ······A············· | ···················· | ···················· | ···················· | ···················· |
| AB | 1911 | hCoV-19/USA/WY-WPHL-20160989/2020\|EPI_ISL_765965\|2 | ···················· | ···················· | ···················· | ···················· | ···················· | ···················· |
| AB | 1912 | hCoV-19/USA/WY-WPHL-20160993/2020\|EPI_ISL_765966\|2 | ···················· | ···················· | ···················· | ···················· | ···················· | ···················· |
| AB | 1913 | hCoV-19/USA/WY-WPHL-20161263/2020\|EPI_ISL_765967\|2 | ···················· | ···················· | ···················· | ···················· | ···················· | ···················· |
| AB | 1914 | hCoV-19/USA/WY-WPHL-20161545/2020\|EPI_ISL_765969\|2 | ···················· | ···················· | ···················· | ···················· | ···················· | ···················· |
| AB | 1915 | hCoV-19/USA/WY-WPHL-20161549/2020\|EPI_ISL_765970\|2 | ···················· | ······A············· | ···················· | ···················· | ···················· | ···················· |
| AB | 1916 | hCoV-19/USA/WY-WPHL-20161554/2020\|EPI_ISL_765971\|2 | ···················· | ···················· | ···················· | ···················· | ···················· | ···················· |
| AB | 1917 | hCoV-19/USA/WY-WPHL-20161708/2020\|EPI_ISL_765972\|2 | ···················· | ···················· | ···················· | ···················· | ···················· | ···················· |
| AB | 1919 | hCoV-19/USA/WY-WPHL-20162190/2020\|EPI_ISL_765975\|2 | ···················· | ···················· | ···················· | ···················· | ···················· | ···················· |
| AB | 1920 | hCoV-19/USA/WY-WPHL-20162212/2020\|EPI_ISL_765976\|2 | ···················· | ···················· | ···················· | ···················· | ···················· | ···················· |
| AB | 1921 | hCoV-19/USA/WY-WPHL-20162292/2020\|EPI_ISL_765977\|2 | ···················· | ···················· | ···················· | ···················· | ···················· | ···················· |
| AB | 1922 | hCoV-19/USA/WY-WPHL-20162761/2020\|EPI_ISL_765978\|2 | ···················· | ···················· | ···················· | ···················· | ···················· | ···················· |
| AB | 1923 | hCoV-19/USA/WY-WPHL-20167571/2020\|EPI_ISL_765979\|2 | ···················· | ···················· | ···················· | ···················· | ···················· | ···················· |
| AB | 1924 | hCoV-19/USA/WY-WPHL-20167581/2020\|EPI_ISL_765980\|2 | ···················· | ···················· | ···················· | ···················· | ···················· | ···················· |
| AB | 1925 | hCoV-19/USA/WY-WPHL-20167589/2020\|EPI_ISL_765981\|2 | ···················· | ···················· | ···················· | ···················· | ···················· | ···················· |
| AB | 1926 | hCoV-19/USA/WY-WPHL-20167591/2020\|EPI_ISL_765982\|2 | ···················· | ···················· | ···················· | ···················· | ···················· | ···················· |
| AB | 1927 | hCoV-19/USA/WY-WPHL-20167677/2020\|EPI_ISL_765983\|2 | ···················· | ···················· | ···················· | ···················· | ···················· | ···················· |
| AB | 1928 | hCoV-19/USA/WY-WPHL-20167879/2020\|EPI_ISL_765985\|2 | ········T··········· | ···················· | ···················· | ···················· | ···················· | ···················· |
| AB | 1929 | hCoV-19/USA/MA-MGH-03584/2020\|EPI_ISL_765986\|2020- | ···················· | ···················· | ···················· | ···················· | ···················· | ···················· |
| AB | 1930 | hCoV-19/USA/CA-LACPHL-AF00061/2020\|EPI_ISL_765998\| | ···················· | ···················· | ···················· | ···················· | ···················· | ···················· |
| AB | 1931 | hCoV-19/USA/CA-LACPHL-AF00062/2020\|EPI_ISL_765999\| | ···················· | ···················· | ···················· | ···················· | ···················· | ···················· |
| AB | 1932 | hCoV-19/Norway/5854/2020\|EPI_ISL_775304\|2020-11-11 | ···················· | ···················· | ···················· | ···················· | ···················· | ···················· |
| AB | 1933 | hCoV-19/Norway/6702/2020\|EPI_ISL_775305\|2020-12-04 | ···················· | ·A·················· | ···················· | ···················· | ···················· | ···A················ |
| AB | 1934 | hCoV-19/Norway/3980/2020\|EPI_ISL_775307\|2020-09-28 | ···················· | ···················· | ···················· | ···················· | ···················· | ···················· |
| AB | 1935 | hCoV-19/Norway/3982/2020\|EPI_ISL_775308\|2020-09-28 | ···················· | ···················· | ···················· | ···················· | ···················· | ···················· |
| AB | 1936 | hCoV-19/Norway/4062/2020\|EPI_ISL_775309\|2020-10-03 | ···················· | ···················· | ···················· | ···················· | ···················· | ···················· |
| AB | 1937 | hCoV-19/Norway/4064/2020\|EPI_ISL_775310\|2020-10-01 | ···················· | ···················· | ···················· | ···················· | ···················· | ···················· |
| AB | 1938 | hCoV-19/Norway/4230/2020\|EPI_ISL_775311\|2020-10-08 | ···················· | ···················· | ···················· | ···················· | ···················· | ···················· |
| AB | 1939 | hCoV-19/Norway/4253/2020\|EPI_ISL_775312\|2020-10-07 | ···················· | ···················· | ···················· | ···················· | ···················· | ···················· |
| AB | 1940 | hCoV-19/Norway/4334/2020\|EPI_ISL_775313\|2020-10-12 | ···················· | ·A·················· | ···················· | ···················· | ···················· | ···················· |
| AB | 1941 | hCoV-19/Norway/4558/2020\|EPI_ISL_775314\|2020-10-13 | ···················· | ···················· | ···················· | ···················· | ···················· | ···················· |
| AB | 1942 | hCoV-19/Norway/4634/2020\|EPI_ISL_775315\|2020-10-18 | ···················· | ···················· | ···················· | ···················· | ···················· | ···················· |
| AB | 1943 | hCoV-19/Norway/4691/2020\|EPI_ISL_775316\|2020-10-07 | ···················· | ···················· | ···················· | ···················· | ···················· | ···················· |
| AB | 1944 | hCoV-19/Norway/4692/2020\|EPI_ISL_775317\|2020-10-05 | ···················· | ···················· | ···················· | ···················· | ···················· | ···················· |
| AB | 1945 | hCoV-19/Norway/4694/2020\|EPI_ISL_775318\|2020-10-07 | ···················· | ·A·················· | ···················· | ···················· | ···················· | ···A················ |
| AB | 1946 | hCoV-19/Norway/4753/2020\|EPI_ISL_775319\|2020-10-20 | ···················· | ···················· | ···················· | ···················· | ···················· | ···················· |
| AB | 1947 | hCoV-19/Norway/4757/2020\|EPI_ISL_775320\|2020-10-19 | ···················· | ···················· | ···················· | ···················· | ···················· | ···················· |
| AB | 1948 | hCoV-19/Norway/4881/2020\|EPI_ISL_775321\|2020-10-25 | ···················· | ···················· | ···················· | ···················· | ···················· | ···················· |
| AB | 1949 | hCoV-19/Norway/5375/2020\|EPI_ISL_775322\|2020-10-10 | ···················· | ···················· | ···················· | ···················· | ···················· | ···················· |
| AB | 1950 | hCoV-19/Norway/5385/2020\|EPI_ISL_775323\|2020-10-27 | ···················· | ···················· | ···················· | ···················· | ···················· | ···················· |
| AB | 1951 | hCoV-19/Norway/5389/2020\|EPI_ISL_775324\|2020-10-22 | ···················· | ·A·················· | ···················· | ···················· | ···················· | ···A················ |
| AB | 1952 | hCoV-19/Norway/5390/2020\|EPI_ISL_775325\|2020-10-19 | ···················· | ·A·················· | ···················· | ···················· | ···················· | ···················· |
| AB | 1953 | hCoV-19/Norway/5391/2020\|EPI_ISL_775326\|2020-10-20 | ···················· | ···················· | ···················· | ···················· | ···················· | ···················· |
| AB | 1954 | hCoV-19/Norway/5411/2020\|EPI_ISL_775327\|2020-10-30 | ···················· | ···················· | ···················· | ···················· | ···················· | ···················· |
| AB | 1955 | hCoV-19/Norway/5412/2020\|EPI_ISL_775328\|2020-11-03 | ···················· | ···················· | ···················· | ···················· | ···················· | ···················· |
| AB | 1956 | hCoV-19/Norway/5413/2020\|EPI_ISL_775329\|2020-11-03 | ···················· | ···················· | ···················· | ···················· | ···················· | ···················· |
| AB | 1957 | hCoV-19/Norway/5465/2020\|EPI_ISL_775330\|2020-10-31 | ···················· | ···················· | ···················· | ···················· | ···················· | ···················· |
| AB | 1958 | hCoV-19/Norway/5754/2020\|EPI_ISL_775331\|2020-10-28 | ···················· | ···················· | ···················· | ···················· | ···················· | ···················· |
| AB | 1959 | hCoV-19/Norway/5759/2020\|EPI_ISL_775332\|2020-10-23 | ···················· | ···················· | ···················· | ···················· | ···················· | ···················· |
| AB | 1960 | hCoV-19/Norway/5795/2020\|EPI_ISL_775333\|2020-11-10 | ···················· | ···················· | ···················· | ···················· | ···················· | ···················· |
| AB | 1961 | hCoV-19/Norway/5830/2020\|EPI_ISL_775334\|2020-11-05 | ···················· | ·A·················· | ···················· | ···················· | ···················· | ···················· |
| AB | 1962 | hCoV-19/Norway/5849/2020\|EPI_ISL_775335\|2020-11-18 | ···················· | ···················· | ···················· | ···················· | ···················· | ···················· |
| AB | 1963 | hCoV-19/Norway/5852/2020\|EPI_ISL_775336\|2020-11-16 | ···················· | ···················· | ···················· | ···················· | ···················· | ···················· |
| AB | 1964 | hCoV-19/Norway/5856/2020\|EPI_ISL_775337\|2020-11-15 | ···················· | ···················· | ···················· | ···················· | ···················· | ···················· |
| AB | 1965 | hCoV-19/Norway/5857/2020\|EPI_ISL_775338\|2020-11-15 | ···················· | ·A·················· | ···················· | ···················· | ···················· | ···A················ |

| | | | | | | | |
|---|---|---|---|---|---|---|---|
| AB | 1966 hCoV-19/Norway/5858/2020\|EPI_ISL_775339\|2020-11-15 | ···················· | ···················· | ···················· | ···················· | ···················· | ···················· |
| AB | 1967 hCoV-19/Norway/5859/2020\|EPI_ISL_775340\|2020-11-14 | ···················· | ···················· | ···················· | ···················· | ···················· | ···················· |
| AB | 1968 hCoV-19/Norway/5860/2020\|EPI_ISL_775341\|2020-11-10 | ···················· | ···················· | ···················· | ···················· | ······T············· | ···················· |
| AB | 1969 hCoV-19/Norway/5861/2020\|EPI_ISL_775342\|2020-11-09 | ···················· | ···················· | ···················· | ···················· | ···················· | ···················· |
| AB | 1970 hCoV-19/Norway/5862/2020\|EPI_ISL_775343\|2020-11-08 | ···················· | ···················· | ···················· | ···················· | ···················· | ···················· |
| AB | 1971 hCoV-19/Norway/5863/2020\|EPI_ISL_775344\|2020-10-28 | ···················· | ···················· | ···················· | ···················· | ···················· | ···················· |
| AB | 1972 hCoV-19/Norway/5877/2020\|EPI_ISL_775345\|2020-11-10 | ···················· | ···················· | ···················· | ···················· | ···················· | ···················· |
| AB | 1973 hCoV-19/Norway/5885/2020\|EPI_ISL_775346\|2020-11-13 | ···················· | ···················· | ···················· | ···················· | ···················· | ···················· |
| AB | 1974 hCoV-19/Norway/5886/2020\|EPI_ISL_775347\|2020-11-14 | ···················· | ···················· | ···················· | ···················· | ···················· | ···················· |
| AB | 1975 hCoV-19/Norway/5900/2020\|EPI_ISL_775348\|2020-11-22 | ···················· | ·A·················· | ···················· | ···················· | ···················· | ···A················ |
| AB | 1976 hCoV-19/Norway/5951/2020\|EPI_ISL_775349\|2020-11-16 | ···················· | ···················· | ···················· | ···················· | ···················· | ···················· |
| AB | 1977 hCoV-19/Norway/5952/2020\|EPI_ISL_775350\|2020-11-17 | ···················· | ···················· | ···················· | ···················· | ···················· | ···················· |
| AB | 1978 hCoV-19/Norway/5953/2020\|EPI_ISL_775351\|2020-11-17 | ···················· | ···················· | ···················· | ···················· | ···················· | ···················· |
| AB | 1979 hCoV-19/Norway/5955/2020\|EPI_ISL_775352\|2020-11-17 | ···················· | ···················· | ···················· | ···················· | ···················· | ···················· |
| AB | 1980 hCoV-19/Norway/6007/2020\|EPI_ISL_775353\|2020-11-15 | ···················· | ···················· | ···················· | ···················· | ······T············· | ···················· |
| AB | 1981 hCoV-19/Norway/6008/2020\|EPI_ISL_775354\|2020-11-16 | ···················· | ···················· | ···················· | ···················· | ······T············· | ···················· |
| AB | 1982 hCoV-19/Norway/6010/2020\|EPI_ISL_775355\|2020-11-15 | ···················· | ···················· | ···················· | ···················· | ······T············· | ···················· |
| AB | 1983 hCoV-19/Norway/6016/2020\|EPI_ISL_775356\|2020-11-26 | ···················· | ···················· | ···················· | ···················· | ···················· | ···················· |
| AB | 1984 hCoV-19/Norway/6235/2020\|EPI_ISL_775357\|2020-11-25 | ···················· | ···················· | ···················· | ···················· | ···················· | ···················· |
| AB | 1985 hCoV-19/Norway/6239/2020\|EPI_ISL_775358\|2020-11-23 | ···················· | ···················· | ···················· | ···················· | ···················· | ···················· |
| AB | 1986 hCoV-19/Israel/CVL-30-ngs/2020\|EPI_ISL_776612\|2020 | ···················· | ···················· | ···················· | ···················· | ···················· | ···················· |
| AB | 1987 hCoV-19/Israel/CVL-36-ngs/2020\|EPI_ISL_776613\|2020 | ···················· | ···················· | ···················· | ···················· | ···················· | ···················· |
| AB | 1988 hCoV-19/Israel/CVL-4-ngs/2020\|EPI_ISL_776614\|2020- | ···················· | ···················· | ···················· | ···················· | ···················· | ···················· |
| AB | 1989 hCoV-19/Israel/CVL-44-ngs/2020\|EPI_ISL_776615\|2020 | ···················· | ···················· | ···················· | ···················· | ···················· | ···················· |
| AB | 1990 hCoV-19/Israel/CVL-52-ngs/2020\|EPI_ISL_776616\|2020 | ···················· | ···················· | ···················· | ···················· | ···················· | ···················· |
| AB | 1991 hCoV-19/Israel/CVL-12-ngs/2020\|EPI_ISL_776621\|2020 | ···················· | ·A·················· | ···················· | ···················· | ···················· | ···················· |
| AB | 1992 hCoV-19/Israel/CVL-13-ngs/2020\|EPI_ISL_776622\|2020 | ···················· | ···················· | ···················· | ···················· | ···················· | ···················· |
| AB | 1993 hCoV-19/Israel/CVL-17-ngs/2020\|EPI_ISL_776623\|2020 | ···················· | ·A·················· | ···················· | ···················· | ···················· | ···················· |
| AB | 1994 hCoV-19/Israel/CVL-18-ngs/2020\|EPI_ISL_776624\|2020 | ···················· | ···················· | ···················· | ···················· | ···················· | ···················· |
| AB | 1995 hCoV-19/Israel/CVL-19-ngs/2020\|EPI_ISL_776625\|2020 | ···················· | ·A·················· | ···················· | ···················· | ···················· | ···················· |
| AB | 1996 hCoV-19/Israel/CVL-27-ngs/2020\|EPI_ISL_776626\|2020 | ···················· | ·A·················· | ···················· | ···················· | ···················· | ···················· |
| AB | 1997 hCoV-19/Israel/CVL-28-ngs/2020\|EPI_ISL_776627\|2020 | ···················· | ···················· | ···················· | ···················· | ···················· | ···················· |
| AB | 1998 hCoV-19/Israel/CVL-34-ngs/2020\|EPI_ISL_776628\|2020 | ···················· | ···················· | ···················· | ···················· | ···················· | ···················· |
| AB | 2001 hCoV-19/Israel/CVL-51-ngs/2020\|EPI_ISL_776636\|2020 | ···················· | ····T··············· | ···················· | ···················· | ···················· | ···················· |
| AB | 2002 hCoV-19/Israel/CVL-54-ngs/2020\|EPI_ISL_776637\|2020 | ···················· | ···················· | ···················· | ···················· | ···················· | ···················· |
| AB | 2003 hCoV-19/Israel/CVL-59_2-ngs/2020\|EPI_ISL_776638\|20 | ···················· | ···················· | ···················· | ···················· | ···················· | ···················· |
| AB | 2004 hCoV-19/Israel/CVL-59_1-ngs/2020\|EPI_ISL_776639\|20 | ···················· | ···················· | ···················· | ···················· | ···················· | ···················· |
| AB | 2005 hCoV-19/Israel/CVL-6-ngs/2020\|EPI_ISL_776640\|2020- | ···················· | ···················· | ···················· | ···················· | ···················· | ···················· |
| AB | 2006 hCoV-19/Israel/CVL-60-ngs/2020\|EPI_ISL_776641\|2020 | ···················· | ·········A·········· | ···················· | ···················· | ···················· | ···················· |
| AB | 2007 hCoV-19/Israel/CVL-62-ngs/2020\|EPI_ISL_776642\|2020 | ···················· | ···················· | ···················· | ···················· | ···················· | ···················· |
| AB | 2008 hCoV-19/Israel/CVL-68-ngs/2020\|EPI_ISL_776643\|2020 | ···················· | ···················· | ···················· | ···················· | ···················· | ···················· |
| AB | 2009 hCoV-19/Israel/CVL-70-ngs/2020\|EPI_ISL_776644\|2020 | ···················· | ·A·················· | ···················· | ···················· | ···················· | ···················· |
| AB | 2010 hCoV-19/Israel/CVL-72-ngs/2020\|EPI_ISL_776645\|2020 | ···················· | ···················· | ···················· | ···················· | ·········T·········· | ···················· |
| AB | 2012 hCoV-19/Israel/CVL-74-ngs/2020\|EPI_ISL_776647\|2020 | ···················· | ···················· | ···················· | ···················· | ···················· | ···················· |
| AB | 2014 hCoV-19/Israel/CVL-77-ngs/2020\|EPI_ISL_776649\|2020 | ···················· | ···················· | ···················· | ···················· | ···················· | ···················· |
| AB | 2015 hCoV-19/Israel/CVL-80-ngs/2020\|EPI_ISL_776650\|2020 | ···················· | ···················· | ···················· | ···················· | ···················· | ···················· |
| AB | 2016 hCoV-19/Israel/CVL-82-ngs/2020\|EPI_ISL_776651\|2020 | ···················· | ···················· | ···················· | ···················· | ·N·················· | ···················· |
| AB | 2017 hCoV-19/Israel/CVL-83-ngs/2020\|EPI_ISL_776652\|2020 | ···················· | ···················· | ···················· | ···················· | ···················· | ···················· |
| AB | 2018 hCoV-19/Israel/CVL-85-ngs/2020\|EPI_ISL_776653\|2020 | ···················· | ···················· | ···················· | ···················· | ···················· | ···················· |
| AB | 2019 hCoV-19/Israel/CVL-88-ngs/2020\|EPI_ISL_776654\|2020 | ···················· | ···················· | ···················· | ···················· | ···················· | ···················· |
| AB | 2020 hCoV-19/Israel/CVL-89-ngs/2020\|EPI_ISL_776655\|2020 | ···················· | ···················· | ···················· | ···················· | ···················· | ···················· |
| AB | 2021 hCoV-19/Israel/CVL-90-ngs/2020\|EPI_ISL_776656\|2020 | ···················· | ···················· | ···················· | ···················· | ···················· | ···················· |
| AB | 2022 hCoV-19/USA/WA-UW-32120/2020\|EPI_ISL_776661\|2020-1 | ···················· | ···················· | ···················· | ···················· | ···················· | ···················· |
| AB | 2023 hCoV-19/USA/WA-UW-47334/2020\|EPI_ISL_776662\|2020-1 | ···················· | ···················· | ···················· | ···················· | ···················· | ···················· |
| AB | 2024 hCoV-19/USA/WA-UW-10883/2020\|EPI_ISL_776663\|2020-0 | ···················· | ···················· | ···················· | ···················· | ···················· | ···················· |
| AB | 2025 hCoV-19/USA/WA-UW-47378/2020\|EPI_ISL_776667\|2020-1 | ···················· | ···················· | ···················· | ···················· | ···················· | ···················· |
| AB | 2026 hCoV-19/USA/WA-UW-14520/2020\|EPI_ISL_776672\|2020-0 | ···················· | ···················· | ···················· | ···················· | ···················· | ···················· |
| AB | 2027 hCoV-19/USA/WA-UW-16209/2020\|EPI_ISL_776673\|2020-0 | ···················· | ···················· | ···················· | ···················· | ···················· | ···················· |
| AB | 2028 hCoV-19/USA/WA-UW-16246/2020\|EPI_ISL_776674\|2020-0 | ···················· | ···················· | ···················· | ···················· | ···················· | ···················· |
| AB | 2029 hCoV-19/USA/WA-UW-10178/2020\|EPI_ISL_776675\|2020-0 | ···················· | ···················· | ···················· | ···················· | ···················· | ···················· |
| AB | 2030 hCoV-19/USA/WA-UW-10505/2020\|EPI_ISL_776676\|2020-0 | ···················· | ···················· | ···················· | ···················· | ···················· | ···················· |
| AB | 2031 hCoV-19/USA/WA-UW-11591/2020\|EPI_ISL_776677\|2020-0 | ···················· | ···················· | ···················· | ···················· | ···················· | ···················· |
| AB | 2032 hCoV-19/USA/WA-UW-13776/2020\|EPI_ISL_776678\|2020-0 | ···················· | ···················· | ···················· | ···················· | ···················· | ···················· |
| AB | 2033 hCoV-19/USA/WA-UW-13874/2020\|EPI_ISL_776679\|2020-0 | ···················· | ···················· | ···················· | ···················· | ···················· | ···················· |
| AB | 2034 hCoV-19/USA/WA-UW-14353/2020\|EPI_ISL_776680\|2020-0 | ···················· | ···················· | ···················· | ···················· | ···················· | ···················· |
| AB | 2035 hCoV-19/USA/WA-UW-14608/2020\|EPI_ISL_776681\|2020-0 | ···················· | ···················· | ···················· | ···················· | ···················· | ···················· |
| AB | 2036 hCoV-19/USA/WA-UW-14623/2020\|EPI_ISL_776682\|2020-0 | ···················· | ·A·················· | ···················· | ···················· | ···················· | ···················· |
| AB | 2037 hCoV-19/USA/WA-UW-15608/2020\|EPI_ISL_776683\|2020-0 | ···················· | ···················· | ···················· | ···················· | ···················· | ···················· |
| AB | 2038 hCoV-19/USA/WA-UW-15796/2020\|EPI_ISL_776684\|2020-0 | ···················· | ···················· | ···················· | ···················· | ···················· | ···················· |
| AB | 2039 hCoV-19/USA/WA-UW-15821/2020\|EPI_ISL_776685\|2020-0 | ···················· | ···················· | ···················· | ···················· | ···················· | ···················· |
| AB | 2040 hCoV-19/USA/WA-UW-17048/2020\|EPI_ISL_776686\|2020-0 | ···················· | ···················· | ···················· | ···················· | ···················· | ···················· |
| AB | 2041 hCoV-19/USA/WA-UW-17637/2020\|EPI_ISL_776687\|2020-0 | ···················· | ·A·················· | ···················· | ···················· | ···················· | ···················· |
| AB | 2042 hCoV-19/USA/WA-UW-24459/2020\|EPI_ISL_776688\|2020-0 | ···················· | ···················· | ···················· | ···················· | ···················· | ···················· |
| AB | 2043 hCoV-19/USA/WA-UW-25085/2020\|EPI_ISL_776689\|2020-0 | ···················· | ···················· | ···················· | ···················· | ···················· | ···················· |
| AB | 2044 hCoV-19/England/CAMC-CB7594/2020\|EPI_ISL_777935\|20 | ···················· | ···················· | ···················· | ···················· | ···················· | ···················· |
| AB | 2045 hCoV-19/England/CAMC-CB8C07/2020\|EPI_ISL_777938\|20 | ···················· | ···················· | ···················· | ···················· | ······T············· | ···················· |
| AB | 2046 hCoV-19/England/CAMC-CB7C35/2020\|EPI_ISL_777942\|20 | ···················· | ···················· | ···················· | ···················· | ···················· | ···················· |
| AB | 2047 hCoV-19/England/CAMC-CB7BFC/2020\|EPI_ISL_777966\|20 | ···················· | ···················· | ···················· | ···················· | ···················· | ···················· |
| AB | 2048 hCoV-19/England/CAMC-CB87BB/2020\|EPI_ISL_777972\|20 | ···················· | ···················· | ···················· | ···················· | ···················· | ···················· |
| AB | 2049 hCoV-19/England/QEUH-C9332B/2020\|EPI_ISL_762003\|20 | ···················· | ···················· | ···················· | ···················· | ···················· | ···················· |

| | | | | | | | |
|---|---|---|---|---|---|---|---|
| AB | 2050 hCoV-19/England/QEUH-C934EC/2020\|EPI_ISL_762051\|20 | ···················· | ···················· | ···················· | ···················· | ···················· | ···················· |
| AB | 2051 hCoV-19/England/QEUH-C92D6A/2020\|EPI_ISL_762055\|20 | ···················· | ···················· | ···················· | ···················· | ···················· | ···················· |
| AB | 2052 hCoV-19/England/CAMC-C995D4/2020\|EPI_ISL_762059\|20 | ···················· | ···················· | ···················· | ···················· | ···················· | ···················· |
| AB | 2053 hCoV-19/England/CAMC-C9901F/2020\|EPI_ISL_762069\|20 | ···················· | ···················· | ···················· | ···················· | ···················· | ···················· |
| AB | 2054 hCoV-19/England/QEUH-C9315E/2020\|EPI_ISL_762080\|20 | ···················· | ···················· | ···················· | ···················· | ···················· | ···················· |
| AB | 2055 hCoV-19/England/CAMC-C994C8/2020\|EPI_ISL_762085\|20 | ···················· | ···················· | ···················· | ···················· | ···················· | ···················· |
| AB | 2056 hCoV-19/England/CAMC-C98B88/2020\|EPI_ISL_762091\|20 | ···················· | ···················· | ···················· | ···················· | ···················· | ···················· |
| AB | 2057 hCoV-19/England/CAMC-C9946E/2020\|EPI_ISL_762097\|20 | ···················· | ···················· | ···················· | ···················· | ······T············· | ···················· |
| AB | 2058 hCoV-19/Italy/UMB-IZSUM-48682-24/2020\|EPI_ISL_7633 | ···················· | ···················· | ···················· | ···················· | ···················· | ···················· |
| AB | 2059 hCoV-19/Italy/UMB-IZSUM-48682-23/2020\|EPI_ISL_7633 | ···················· | ···················· | ···················· | ···················· | ···················· | ···················· |
| AB | 2060 hCoV-19/Italy/UMB-IZSUM-48682-22/2020\|EPI_ISL_7633 | ···················· | ···················· | ···················· | ···················· | ···················· | ···················· |
| AB | 2061 hCoV-19/Italy/UMB-IZSUM-48682-21/2020\|EPI_ISL_7633 | ···················· | ···················· | ···················· | ···················· | ···················· | ···················· |
| AB | 2062 hCoV-19/Italy/UMB-IZSUM-48682-8/2020\|EPI_ISL_76332 | ···················· | ···················· | ···················· | ···················· | ······T············· | ···················· |
| AB | 2063 hCoV-19/Italy/UMB-IZSUM-48682-2/2020\|EPI_ISL_76333 | ···················· | ···················· | ···················· | ···················· | ······T············· | ···················· |
| AB | 2064 hCoV-19/Wales/PHWC-4A4073/2020\|EPI_ISL_764656\|2020 | ···················· | ···················· | ···················· | ···················· | ···················· | ···················· |
| AB | 2065 hCoV-19/Wales/PHWC-4A4134/2020\|EPI_ISL_764657\|2020 | ···················· | ···················· | ···················· | ···················· | ···················· | ···················· |
| AB | 2068 hCoV-19/Wales/PHWC-4A419E/2020\|EPI_ISL_764660\|2020 | ···················· | ···················· | ···················· | ···················· | ···················· | ···················· |
| AB | 2074 hCoV-19/Wales/PHWC-4A4204/2020\|EPI_ISL_764666\|2020 | ···················· | ···················· | ···················· | ···················· | ···················· | ···················· |
| AB | 2077 hCoV-19/Wales/PHWC-4A44FF/2020\|EPI_ISL_764702\|2020 | ···················· | ···················· | ···················· | ···················· | ···················· | ···················· |
| AB | 2079 hCoV-19/Wales/PHWC-4A4529/2020\|EPI_ISL_764704\|2020 | ···················· | ···················· | ···················· | ···················· | ···················· | ···················· |
| AB | 2082 hCoV-19/Wales/PHWC-4A4574/2020\|EPI_ISL_764707\|2020 | ···················· | ···················· | ···················· | ···················· | ···················· | ···················· |
| AB | 2084 hCoV-19/Wales/PHWC-4A45A1/2020\|EPI_ISL_764709\|2020 | ···················· | ···················· | ···················· | ···················· | ···················· | ···················· |
| AB | 2085 hCoV-19/Wales/PHWC-4A45B0/2020\|EPI_ISL_764710\|2020 | ···················· | ···················· | ···················· | ···················· | ···················· | ···················· |
| AB | 2088 hCoV-19/USA/CA-LACPHL-AF00039/2020\|EPI_ISL_765987\| | ···················· | ···················· | ···················· | ···················· | ···················· | ···················· |
| AB | 2089 hCoV-19/USA/CA-LACPHL-AF00040/2020\|EPI_ISL_765988\| | ···················· | ···················· | ···················· | ···················· | ···················· | ···················· |
| AB | 2090 hCoV-19/USA/CA-LACPHL-AF00041/2020\|EPI_ISL_765989\| | ···················· | ···················· | ···················· | ···················· | ···················· | ···················· |
| AB | 2091 hCoV-19/USA/CA-LACPHL-AF00043/2020\|EPI_ISL_765990\| | ···················· | ···················· | ···················· | ···················· | ···················· | ···················· |
| AB | 2092 hCoV-19/USA/CA-LACPHL-AF00044/2020\|EPI_ISL_765991\| | ···················· | ···················· | ···················· | ···················· | ···················· | ···················· |
| AB | 2093 hCoV-19/USA/CA-LACPHL-AF00045/2020\|EPI_ISL_765992\| | ···················· | ···················· | ···················· | ···················· | ···················· | ···················· |
| AB | 2094 hCoV-19/USA/CA-LACPHL-AF00046/2020\|EPI_ISL_765993\| | ···················· | ···················· | ···················· | ···················· | ···················· | ···················· |
| AB | 2095 hCoV-19/USA/CA-LACPHL-AE00057/2020\|EPI_ISL_765995\| | ···················· | ····T··············· | ···················· | ···················· | ···················· | ···················· |
| AB | 2096 hCoV-19/USA/CA-LACPHL-AE00059/2020\|EPI_ISL_765997\| | ···················· | ···················· | ···················· | ···················· | ···················· | ···················· |
| AB | 2098 hCoV-19/Switzerland/SO-ETHZ-420722/2020\|EPI_ISL_76 | ···················· | ···················· | ···················· | ···················· | ···················· | ···················· |
| AB | 2099 hCoV-19/Switzerland/SO-ETHZ-420326/2020\|EPI_ISL_76 | ···················· | ···················· | ···················· | ···················· | ···················· | ···················· |
| AB | 2100 hCoV-19/Switzerland/SO-ETHZ-420715/2020\|EPI_ISL_76 | ···················· | ···················· | ···················· | ···················· | ···················· | ···················· |
| AB | 2101 hCoV-19/Switzerland/SO-ETHZ-420476/2020\|EPI_ISL_76 | ···················· | ···················· | ···················· | ···················· | ···················· | ···················· |
| AB | 2106 hCoV-19/Switzerland/ZH-ETHZ-420608/2020\|EPI_ISL_76 | ···················· | ···················· | ···················· | ···················· | ···················· | ···················· |
| AB | 2107 hCoV-19/Switzerland/ZH-ETHZ-420661/2020\|EPI_ISL_76 | ···················· | ·A·················· | ···················· | ···················· | ·······S············ | ···A·M·············· |
| AB | 2108 hCoV-19/Switzerland/ZH-ETHZ-420723/2020\|EPI_ISL_76 | ···················· | ···················· | ···················· | ···················· | ···················· | ···················· |
| AB | 2109 hCoV-19/England/QEUH-C5B8F3/2020\|EPI_ISL_769503\|20 | ···················· | ···················· | ···················· | ···················· | ···················· | ···················· |
| AB | 2110 hCoV-19/England/MILK-C60DB0/2020\|EPI_ISL_769507\|20 | ···················· | ···················· | ···················· | ···················· | ···················· | ···················· |
| AB | 2111 hCoV-19/England/MILK-C60D38/2020\|EPI_ISL_769508\|20 | ···················· | ···················· | ···················· | ···················· | ···················· | ···················· |
| AB | 2112 hCoV-19/England/QEUH-C5CA47/2020\|EPI_ISL_769511\|20 | ···················· | ···················· | ···················· | ···················· | ···················· | ···················· |
| AB | 2113 hCoV-19/England/QEUH-C5BB63/2020\|EPI_ISL_769513\|20 | ···················· | ···················· | ···················· | ···················· | ···················· | ···················· |
| AB | 2114 hCoV-19/England/QEUH-C5D92B/2020\|EPI_ISL_769517\|20 | ···················· | ···················· | ···················· | ···················· | ···················· | ···················· |
| AB | 2115 hCoV-19/England/QEUH-C5CA0B/2020\|EPI_ISL_769519\|20 | ···················· | ···················· | ···················· | ···················· | ···················· | ···················· |
| AB | 2116 hCoV-19/England/MILK-C60FC9/2020\|EPI_ISL_769520\|20 | ···················· | ···················· | ···················· | ···················· | ···················· | ···················· |
| AB | 2117 hCoV-19/England/MILK-C6146F/2020\|EPI_ISL_769523\|20 | ···················· | ···················· | ···················· | ···················· | ···················· | ···················· |
| AB | 2118 hCoV-19/England/QEUH-C5CA74/2020\|EPI_ISL_769524\|20 | ···················· | ···················· | ···················· | ···················· | ···················· | ···················· |
| AB | 2119 hCoV-19/England/QEUH-C5C9D1/2020\|EPI_ISL_769525\|20 | ···················· | ···················· | ···················· | ···················· | ···················· | ···················· |
| AB | 2120 hCoV-19/England/QEUH-C5CAB0/2020\|EPI_ISL_769528\|20 | ···················· | ···················· | ···················· | ···················· | ···················· | ···················· |
| AB | 2121 hCoV-19/England/MILK-C60C1D/2020\|EPI_ISL_769529\|20 | ···················· | ···················· | ···················· | ···················· | ···················· | ···················· |
| AB | 2122 hCoV-19/England/QEUH-C5C42B/2020\|EPI_ISL_769530\|20 | ···················· | ···················· | ···················· | ···················· | ···················· | ···················· |
| AB | 2123 hCoV-19/England/MILK-C5FFCD/2020\|EPI_ISL_769531\|20 | ···················· | ···················· | ···················· | ···················· | ···················· | ···················· |
| AB | 2124 hCoV-19/England/QEUH-C5CACF/2020\|EPI_ISL_769533\|20 | ···················· | ···················· | ···················· | ···················· | ···················· | ···················· |
| AB | 2125 hCoV-19/England/MILK-C632DC/2020\|EPI_ISL_769534\|20 | ···················· | ···················· | ···················· | ···················· | ···················· | ···················· |
| AB | 2126 hCoV-19/England/MILK-C60C77/2020\|EPI_ISL_769536\|20 | ···················· | ···················· | ···················· | ···················· | ···················· | ···················· |
| AB | 2127 hCoV-19/England/MILK-C60CD1/2020\|EPI_ISL_769537\|20 | ···················· | ···················· | ···················· | ···················· | ···················· | ···················· |
| AB | 2128 hCoV-19/England/QEUH-C5CA29/2020\|EPI_ISL_769540\|20 | ···················· | ···················· | ···················· | ···················· | ···················· | ···················· |
| AB | 2129 hCoV-19/England/MILK-C51963/2020\|EPI_ISL_769544\|20 | ···················· | ···················· | ···················· | ···················· | ······T············· | ···················· |
| AB | 2130 hCoV-19/England/MILK-C517A5/2020\|EPI_ISL_769545\|20 | ···················· | ···················· | ···················· | ···················· | ···················· | ···················· |
| AB | 2131 hCoV-19/England/MILK-C501F3/2020\|EPI_ISL_769546\|20 | ···················· | ···················· | ···················· | ···················· | ···················· | ···················· |
| AB | 2132 hCoV-19/England/MILK-C5347F/2020\|EPI_ISL_769547\|20 | ···················· | ···················· | ···················· | ···················· | ···················· | ···················· |
| AB | 2133 hCoV-19/England/MILK-C5022D/2020\|EPI_ISL_769548\|20 | ···················· | ···················· | ···················· | ···················· | ···················· | ···················· |
| AB | 2134 hCoV-19/England/MILK-C516C6/2020\|EPI_ISL_769550\|20 | ···················· | ···················· | ···················· | ···················· | ···················· | ···················· |
| AB | 2135 hCoV-19/England/QEUH-C4B6DE/2020\|EPI_ISL_769551\|20 | ···················· | ···················· | ···················· | ···················· | ···················· | ···················· |
| AB | 2136 hCoV-19/England/MILK-C4FE3C/2020\|EPI_ISL_769552\|20 | ···················· | ···················· | ···················· | ···················· | ···················· | ···················· |
| AB | 2137 hCoV-19/England/MILK-C5023C/2020\|EPI_ISL_769555\|20 | ···················· | ···················· | ···················· | ···················· | ···················· | ···················· |
| AB | 2138 hCoV-19/England/QEUH-C4B814/2020\|EPI_ISL_769556\|20 | ···················· | ···················· | ···················· | ···················· | ···················· | ···················· |
| AB | 2139 hCoV-19/England/MILK-C50111/2020\|EPI_ISL_769558\|20 | ···················· | ···················· | ···················· | ···················· | ······T············· | ···················· |
| AB | 2140 hCoV-19/England/MILK-C51936/2020\|EPI_ISL_769561\|20 | ···················· | ···················· | ···················· | ···················· | ···················· | ···················· |
| AB | 2141 hCoV-19/England/MILK-C51884/2020\|EPI_ISL_769563\|20 | ···················· | ···················· | ···················· | ···················· | ···················· | ···················· |
| AB | 2142 hCoV-19/England/MILK-C53A22/2020\|EPI_ISL_769564\|20 | ···················· | ···················· | ···················· | ···················· | ···················· | ···················· |
| AB | 2143 hCoV-19/England/QEUH-C4B8D8/2020\|EPI_ISL_769567\|20 | ···················· | ···················· | ···················· | ···················· | ···················· | ···················· |
| AB | 2144 hCoV-19/England/MILK-C5020F/2020\|EPI_ISL_769569\|20 | ···················· | ···················· | ···················· | ···················· | ···················· | ···················· |
| AB | 2145 hCoV-19/England/MILK-C500F6/2020\|EPI_ISL_769574\|20 | ···················· | ···················· | ···················· | ···················· | ······T············· | ···················· |
| AB | 2146 hCoV-19/England/MILK-C518B1/2020\|EPI_ISL_769578\|20 | ···················· | ···················· | ···················· | ···················· | ···················· | ···················· |
| AB | 2147 hCoV-19/England/MILK-C51848/2020\|EPI_ISL_769580\|20 | ···················· | ···················· | ···················· | ···················· | ···················· | ···················· |
| AB | 2148 hCoV-19/England/QEUH-C4BAD2/2020\|EPI_ISL_769583\|20 | ···················· | ···················· | ···················· | ···················· | ······T············· | ···················· |
| AB | 2149 hCoV-19/England/MILK-C51990/2020\|EPI_ISL_769584\|20 | ···················· | ···················· | ···················· | ···················· | ···················· | ···················· |

| | | | | | | | |
|---|---|---|---|---|---|---|---|
| AB | 2150 hCoV-19/England/MILK-C518A2/2020\|EPI_ISL_769585\|20 | ···················· | ···················· | ···················· | ···················· | ···················· | ···················· |
| AB | 2151 hCoV-19/England/MILK-C533FA/2020\|EPI_ISL_769586\|20 | ···················· | ···················· | ···················· | ···················· | ···················· | ···················· |
| AB | 2152 hCoV-19/England/MILK-C50278/2020\|EPI_ISL_769587\|20 | ···················· | ···················· | ···················· | ···················· | ···················· | ···················· |
| AB | 2153 hCoV-19/England/MILK-C4FDF3/2020\|EPI_ISL_769591\|20 | ···················· | ···················· | ···················· | ···················· | ···················· | ···················· |
| AB | 2154 hCoV-19/England/MILK-C51981/2020\|EPI_ISL_769592\|20 | ···················· | ···················· | ···················· | ···················· | ···················· | ···················· |
| AB | 2155 hCoV-19/England/QEUH-C4BBEE/2020\|EPI_ISL_769594\|20 | ············T······· | ···················· | ···················· | ···················· | ···················· | ···················· |
| AB | 2156 hCoV-19/England/MILK-C4FE5A/2020\|EPI_ISL_769596\|20 | ···················· | ···················· | ···················· | ···················· | ···················· | ···················· |
| AB | 2157 hCoV-19/England/MILK-C517B4/2020\|EPI_ISL_769597\|20 | ···················· | ···················· | ···················· | ···················· | ···················· | ···················· |
| AB | 2158 hCoV-19/Lithuania/MR-LUHS-Eilnr234964/2020\|EPI_ISL | ···················· | ···················· | ···················· | ···················· | ···················· | ···················· |
| AB | 2159 hCoV-19/Lithuania/MR-LUHS-Eilnr235011/2020\|EPI_ISL | ···················· | ···················· | ···················· | ···················· | ···················· | ···················· |
| AB | 2160 hCoV-19/Lithuania/MR-LUHS-Eilnr235073/2020\|EPI_ISL | ···················· | ···················· | ···················· | ···················· | ···················· | ···················· |
| AB | 2161 hCoV-19/Lithuania/MR-LUHS-Eilnr235793/2020\|EPI_ISL | ···················· | ···················· | ···················· | ···················· | ···················· | ···················· |
| AB | 2162 hCoV-19/Lithuania/MR-LUHS-Eilnr235905/2020\|EPI_ISL | ···················· | ···················· | ···················· | ···················· | ···················· | ···················· |
| AB | 2163 hCoV-19/Lithuania/MR-LUHS-Eilnr236244/2020\|EPI_ISL | ···················· | ···················· | ···················· | ···················· | ···················· | ···················· |
| AB | 2164 hCoV-19/Lithuania/MR-LUHS-Eilnr236247/2020\|EPI_ISL | ···················· | ···················· | ···················· | ···················· | ···················· | ···················· |
| AB | 2165 hCoV-19/Lithuania/MR-LUHS-Eilnr236309/2020\|EPI_ISL | ···················· | ···················· | ···················· | ···················· | ···················· | ···················· |
| AB | 2166 hCoV-19/Lithuania/MR-LUHS-Eilnr236563/2020\|EPI_ISL | ···················· | ···················· | ···················· | ···················· | ···················· | ···················· |
| AB | 2167 hCoV-19/Lithuania/MR-LUHS-Eilnr236583/2020\|EPI_ISL | ···················· | ···················· | ···················· | ···················· | ···················· | ···················· |
| AB | 2168 hCoV-19/Lithuania/MR-LUHS-Eilnr236584/2020\|EPI_ISL | ···················· | ···················· | ···················· | ···················· | ···················· | ···················· |
| AB | 2169 hCoV-19/Lithuania/MR-LUHS-Eilnr236600/2020\|EPI_ISL | ···················· | ···················· | ···················· | ···················· | ···················· | ···················· |
| AB | 2170 hCoV-19/Lithuania/MR-LUHS-Eilnr236601/2020\|EPI_ISL | ···················· | ···················· | ···················· | ···················· | ···················· | ···················· |
| AB | 2171 hCoV-19/Lithuania/MR-LUHS-Eilnr236606/2020\|EPI_ISL | ···················· | ···················· | ···················· | ···················· | ···················· | ···················· |
| AB | 2172 hCoV-19/Lithuania/MR-LUHS-Eilnr237105/2020\|EPI_ISL | ···················· | ···················· | ···················· | ···················· | ···················· | ···················· |
| AB | 2173 hCoV-19/Lithuania/MR-LUHS-Eilnr237251/2020\|EPI_ISL | ···················· | ···················· | ···················· | ···················· | ···················· | ···················· |
| AB | 2174 hCoV-19/Lithuania/MR-LUHS-Eilnr237513/2020\|EPI_ISL | ···················· | ···················· | ···················· | ···················· | ···················· | ···················· |
| AB | 2175 hCoV-19/Lithuania/MR-LUHS-Eilnr237525/2020\|EPI_ISL | ···················· | ···················· | ···················· | ···················· | ···················· | ···················· |
| AB | 2176 hCoV-19/Lithuania/MR-LUHS-Eilnr237556/2020\|EPI_ISL | ···················· | ···················· | ···················· | ···················· | ···················· | ···················· |
| AB | 2177 hCoV-19/Lithuania/MR-LUHS-Eilnr237653/2020\|EPI_ISL | ···················· | ···················· | ···················· | ···················· | ···················· | ···················· |
| AB | 2178 hCoV-19/Lithuania/MR-LUHS-Eilnr237654/2020\|EPI_ISL | ···················· | ···················· | ···················· | ···················· | ···················· | ···················· |
| AB | 2179 hCoV-19/Lithuania/MR-LUHS-Eilnr238011/2020\|EPI_ISL | ···················· | ···················· | ···················· | ···················· | ···················· | ···················· |
| AB | 2180 hCoV-19/Lithuania/MR-LUHS-Eilnr238179/2020\|EPI_ISL | ···················· | ···················· | ···················· | ···················· | ···················· | ···················· |
| AB | 2181 hCoV-19/Lithuania/MR-LUHS-Eilnr238180/2020\|EPI_ISL | ···················· | ···················· | ···················· | ···················· | ···················· | ···················· |
| AB | 2182 hCoV-19/Lithuania/MR-LUHS-Eilnr238208/2020\|EPI_ISL | ···················· | ···················· | ···················· | ···················· | ···················· | ···················· |
| AB | 2183 hCoV-19/Lithuania/MR-LUHS-Eilnr238236/2020\|EPI_ISL | ···················· | ···················· | ···················· | ···················· | ···················· | ···················· |
| AB | 2184 hCoV-19/Lithuania/MR-LUHS-Eilnr238273/2020\|EPI_ISL | ···················· | ···················· | ···················· | ···················· | ···················· | ···················· |
| AB | 2185 hCoV-19/Lithuania/MR-LUHS-Eilnr238277/2020\|EPI_ISL | ···················· | ···················· | ···················· | ···················· | ···················· | ···················· |
| AB | 2186 hCoV-19/Lithuania/MR-LUHS-Eilnr238309/2020\|EPI_ISL | ···················· | ···················· | ···················· | ···················· | ···················· | ···················· |
| AB | 2187 hCoV-19/Lithuania/MR-LUHS-Eilnr238326/2020\|EPI_ISL | ···················· | ···················· | ···················· | ···················· | ···················· | ···················· |
| AB | 2188 hCoV-19/Lithuania/MR-LUHS-Eilnr238362/2020\|EPI_ISL | ···················· | ···················· | ···················· | ···················· | ···················· | ···················· |
| AB | 2189 hCoV-19/Lithuania/MR-LUHS-Eilnr238395/2020\|EPI_ISL | ···················· | ···················· | ···················· | ···················· | ···················· | ···················· |
| AB | 2190 hCoV-19/Lithuania/MR-LUHS-Eilnr238418/2020\|EPI_ISL | ···················· | ···················· | ···················· | ···················· | ···················· | ···················· |
| AB | 2191 hCoV-19/Lithuania/MR-LUHS-Eilnrj1/2020\|EPI_ISL_770 | ···················· | ···················· | ···················· | ···················· | ···················· | ···················· |
| AB | 2192 hCoV-19/Lithuania/MR-LUHS-Eilnrj12/2020\|EPI_ISL_77 | ···················· | ···················· | ···················· | ···················· | ···················· | ···················· |
| AB | 2193 hCoV-19/Lithuania/MR-LUHS-Eilnrj14/2020\|EPI_ISL_77 | ···················· | ···················· | ···················· | ···················· | ···················· | ···················· |
| AB | 2194 hCoV-19/Lithuania/MR-LUHS-Eilnrj15/2020\|EPI_ISL_77 | ···················· | ···················· | ···················· | ···················· | ···················· | ···················· |
| AB | 2195 hCoV-19/Lithuania/MR-LUHS-Eilnrj17/2020\|EPI_ISL_77 | ···················· | ···················· | ···················· | ···················· | ···················· | ···················· |
| AB | 2196 hCoV-19/Lithuania/MR-LUHS-Eilnrj18/2020\|EPI_ISL_77 | ···················· | ···················· | ···················· | ···················· | ···················· | ···················· |
| AB | 2197 hCoV-19/Lithuania/MR-LUHS-Eilnrj20/2020\|EPI_ISL_77 | ···················· | ···················· | ···················· | ···················· | ···················· | ···················· |
| AB | 2198 hCoV-19/Lithuania/MR-LUHS-Eilnrj21/2020\|EPI_ISL_77 | ···················· | ···················· | ···················· | ···················· | ···················· | ···················· |
| AB | 2199 hCoV-19/Lithuania/MR-LUHS-Eilnrj22/2020\|EPI_ISL_77 | ···················· | ···················· | ···················· | ···················· | ···················· | ···················· |
| AB | 2200 hCoV-19/Lithuania/MR-LUHS-Eilnrj23/2020\|EPI_ISL_77 | ···················· | ···················· | ···················· | ···················· | ···················· | ···················· |
| AB | 2201 hCoV-19/Lithuania/MR-LUHS-Eilnrj24/2020\|EPI_ISL_77 | ···················· | ···················· | ···················· | ···················· | ···················· | ···················· |
| AB | 2202 hCoV-19/Lithuania/MR-LUHS-Eilnrj5/2020\|EPI_ISL_770 | ···················· | ···················· | ···················· | ···················· | ···················· | ···················· |
| AB | 2203 hCoV-19/Lithuania/MR-LUHS-Eilnrj6/2020\|EPI_ISL_770 | ···················· | ···················· | ···················· | ···················· | ···················· | ···················· |
| AB | 2204 hCoV-19/Lithuania/MR-LUHS-Eilnrj8/2020\|EPI_ISL_770 | ···················· | ···················· | ···················· | ···················· | ···················· | ···················· |
| AB | 2205 hCoV-19/Lithuania/MR-LUHS-Eilnrj9/2020\|EPI_ISL_770 | ···················· | ···················· | ···················· | ···················· | ···················· | ···················· |
| AB | 2206 hCoV-19/Lithuania/MR-LUHS-Eilnr569/2020\|EPI_ISL_77 | ···················· | ···················· | ···················· | ···················· | ···················· | ···················· |
| AB | 2207 hCoV-19/Lithuania/MR-LUHS-Eilnr507/2020\|EPI_ISL_77 | ···················· | ···················· | ···················· | ···················· | ···················· | ···················· |
| AB | 2208 hCoV-19/Brazil/RS-00651/2020\|EPI_ISL_770551\|2020-1 | ···················· | ···················· | ···················· | ···················· | ···················· | ···················· |
| AB | 2209 hCoV-19/Brazil/RS-00655/2020\|EPI_ISL_770552\|2020-1 | ···················· | ···················· | ···················· | ···················· | ···················· | ···················· |
| AB | 2210 hCoV-19/Brazil/RS-00628/2020\|EPI_ISL_770553\|2020-1 | ···················· | C··················· | ···················· | ···················· | ···················· | ···················· |
| AB | 2211 hCoV-19/Brazil/RS-00682/2020\|EPI_ISL_770554\|2020-1 | ···················· | C··················· | ···················· | ···················· | ···················· | ···················· |
| AB | 2212 hCoV-19/Brazil/RS-00603/2020\|EPI_ISL_770555\|2020-1 | ···················· | ···················· | ···················· | ···················· | ···················· | ···················· |
| AB | 2213 hCoV-19/Brazil/RS-00604/2020\|EPI_ISL_770556\|2020-1 | ···················· | ···················· | ···················· | ···················· | ···················· | ···················· |
| AB | 2214 hCoV-19/Brazil/RS-00605/2020\|EPI_ISL_770557\|2020-1 | ···················· | ···················· | ···················· | ···················· | ···················· | ···················· |
| AB | 2215 hCoV-19/Brazil/RS-00606/2020\|EPI_ISL_770558\|2020-1 | ···················· | ···················· | ···················· | ···················· | ···················· | ···················· |
| AB | 2216 hCoV-19/Brazil/RS-00607/2020\|EPI_ISL_770559\|2020-1 | ···················· | ···················· | ···················· | ···················· | ···················· | ···················· |
| AB | 2217 hCoV-19/Brazil/RS-00608/2020\|EPI_ISL_770560\|2020-1 | ···················· | ···················· | ···················· | ···················· | ···················· | ···················· |
| AB | 2218 hCoV-19/Brazil/RS-00609/2020\|EPI_ISL_770561\|2020-1 | ···················· | ···················· | ···················· | ···················· | ···················· | ···················· |
| AB | 2219 hCoV-19/Brazil/RS-00610/2020\|EPI_ISL_770562\|2020-1 | ···················· | ···················· | ···················· | ···················· | ···················· | ···················· |
| AB | 2220 hCoV-19/Brazil/RS-00611/2020\|EPI_ISL_770563\|2020-1 | ···················· | ···················· | ···················· | ···················· | ···················· | ···················· |
| AB | 2221 hCoV-19/Brazil/RS-00612/2020\|EPI_ISL_770564\|2020-1 | ···················· | ···················· | ···················· | ···················· | ···················· | ···················· |
| AB | 2222 hCoV-19/Brazil/RS-00613/2020\|EPI_ISL_770565\|2020-1 | ···················· | ···················· | ···················· | ···················· | ···················· | ···················· |
| AB | 2223 hCoV-19/Brazil/RS-00614/2020\|EPI_ISL_770566\|2020-1 | ···················· | ···················· | ···················· | ···················· | ···················· | ···················· |
| AB | 2224 hCoV-19/Brazil/RS-00615/2020\|EPI_ISL_770567\|2020-1 | ···················· | ···················· | ···················· | ···················· | ···················· | ···················· |
| AB | 2225 hCoV-19/Brazil/RS-00616/2020\|EPI_ISL_770568\|2020-1 | ···················· | ···················· | ···················· | ···················· | ···················· | ···················· |
| AB | 2226 hCoV-19/Brazil/RS-00617/2020\|EPI_ISL_770569\|2020-1 | ···················· | ···················· | ···················· | ···················· | ···················· | ···················· |
| AB | 2227 hCoV-19/Brazil/RS-00618/2020\|EPI_ISL_770570\|2020-1 | ···················· | ···················· | ···················· | ···················· | ···················· | ···················· |
| AB | 2228 hCoV-19/Brazil/RS-00619/2020\|EPI_ISL_770571\|2020-1 | ···················· | ···················· | ···················· | ···················· | ···················· | ···················· |
| AB | 2229 hCoV-19/Brazil/RS-00620/2020\|EPI_ISL_770572\|2020-1 | ···················· | ···················· | ···················· | ···················· | ···················· | ···················· |

| | | | | | | | | |
|---|---|---|---|---|---|---|---|---|
| AB | 2230 | hCoV-19/Brazil/RS-00622/2020\|EPI_ISL_770573\|2020-1 | ···················· | ···················· | ···················· | ···················· | ···················· | ···················· |
| AB | 2231 | hCoV-19/Brazil/RS-00623/2020\|EPI_ISL_770574\|2020-1 | ···················· | ···················· | ···················· | ···················· | ···················· | ···················· |
| AB | 2232 | hCoV-19/Brazil/RS-00625/2020\|EPI_ISL_770575\|2020-1 | ···················· | ···················· | ···················· | ···················· | ···················· | ···················· |
| AB | 2233 | hCoV-19/Brazil/RS-00626/2020\|EPI_ISL_770576\|2020-1 | ···················· | ···················· | ···················· | ···················· | ···················· | ···················· |
| AB | 2234 | hCoV-19/Brazil/RS-00627/2020\|EPI_ISL_770577\|2020-1 | ···················· | ···················· | ···················· | ···················· | ···················· | ···················· |
| AB | 2235 | hCoV-19/Brazil/RS-00629/2020\|EPI_ISL_770578\|2020-1 | ···················· | ···················· | ···················· | ···················· | ···················· | ···················· |
| AB | 2236 | hCoV-19/Brazil/RS-00630/2020\|EPI_ISL_770579\|2020-1 | ···················· | ···················· | ···················· | ···················· | ···················· | ···················· |
| AB | 2237 | hCoV-19/Brazil/RS-00632/2020\|EPI_ISL_770580\|2020-1 | ···················· | ···················· | ···················· | ···················· | ···················· | ···················· |
| AB | 2238 | hCoV-19/Brazil/RS-00633/2020\|EPI_ISL_770581\|2020-1 | ···················· | ···················· | ···················· | ···················· | ···················· | ···················· |
| AB | 2239 | hCoV-19/Brazil/RS-00634/2020\|EPI_ISL_770582\|2020-1 | ···················· | ···················· | ···················· | ···················· | ···················· | ···················· |
| AB | 2240 | hCoV-19/Brazil/RS-00635/2020\|EPI_ISL_770583\|2020-1 | ···················· | ···················· | ···················· | ···················· | ···················· | ···················· |
| AB | 2241 | hCoV-19/Brazil/RS-00636/2020\|EPI_ISL_770584\|2020-1 | ···················· | ···················· | ···················· | ···················· | ···················· | ···················· |
| AB | 2242 | hCoV-19/Norway/5841/2020\|EPI_ISL_775300\|2020-11-13 | ···················· | ···················· | ···················· | ···················· | ···················· | ···················· |
| AB | 2243 | hCoV-19/Norway/5842/2020\|EPI_ISL_775301\|2020-11-13 | ···················· | ···················· | ···················· | ···················· | ···················· | ···················· |
| AB | 2244 | hCoV-19/Norway/5846/2020\|EPI_ISL_775302\|2020-11-11 | ···················· | ···················· | ···················· | ···················· | ···················· | ···················· |
| AB | 2245 | hCoV-19/Norway/5847/2020\|EPI_ISL_775303\|2020-11-13 | ···················· | ···················· | ···················· | ···················· | ···················· | ···················· |
| AB | 2246 | hCoV-19/Norway/6241/2020\|EPI_ISL_775359\|2020-11-23 | ···················· | ···················· | ···················· | ···················· | ···················· | ···················· |
| AB | 2247 | hCoV-19/Norway/6242/2020\|EPI_ISL_775360\|2020-11-23 | ···················· | ·A·················· | ···················· | ···················· | ···················· | ···A················ |
| AB | 2248 | hCoV-19/Norway/6243/2020\|EPI_ISL_775361\|2020-11-23 | ···················· | ···················· | ···················· | ···················· | ···················· | ···················· |
| AB | 2249 | hCoV-19/Norway/6244/2020\|EPI_ISL_775362\|2020-11-24 | ···················· | ···················· | ···················· | ···················· | ······T············· | ···················· |
| AB | 2250 | hCoV-19/Norway/6245/2020\|EPI_ISL_775363\|2020-11-18 | ···················· | ···················· | ···················· | ···················· | ······T············· | ···················· |
| AB | 2251 | hCoV-19/Norway/6246/2020\|EPI_ISL_775364\|2020-11-16 | ···················· | ···················· | ···················· | ···················· | ···················· | ···················· |
| AB | 2252 | hCoV-19/Norway/6247/2020\|EPI_ISL_775365\|2020-11-16 | ···················· | ···················· | ···················· | ···················· | ···················· | ···················· |
| AB | 2253 | hCoV-19/Norway/6248/2020\|EPI_ISL_775366\|2020-11-16 | ···················· | ···················· | ···················· | ···················· | ···················· | ···················· |
| AB | 2254 | hCoV-19/Norway/6249/2020\|EPI_ISL_775367\|2020-11-18 | ···················· | ···················· | ···················· | ···················· | ···················· | ···················· |
| AB | 2255 | hCoV-19/Norway/6251/2020\|EPI_ISL_775368\|2020-11-24 | ···················· | ···················· | ···················· | ···················· | ···················· | ···················· |
| AB | 2256 | hCoV-19/Norway/6252/2020\|EPI_ISL_775369\|2020-11-25 | ···················· | ···················· | ···················· | ···················· | ···················· | ···················· |
| AB | 2257 | hCoV-19/Norway/6253/2020\|EPI_ISL_775370\|2020-11-27 | ···················· | ···················· | ···················· | ···················· | ···················· | ···················· |
| AB | 2258 | hCoV-19/Norway/6265/2020\|EPI_ISL_775371\|2020-11-30 | ···················· | ···················· | ···················· | ···················· | ···················· | ···················· |
| AB | 2259 | hCoV-19/Norway/6268/2020\|EPI_ISL_775372\|2020-10-23 | ···················· | ···················· | ···················· | ···················· | ···················· | ···················· |
| AB | 2260 | hCoV-19/Norway/6275/2020\|EPI_ISL_775373\|2020-11-25 | ···················· | ····A··············· | ···················· | ···················· | ···················· | ···················· |
| AB | 2261 | hCoV-19/Norway/6276/2020\|EPI_ISL_775374\|2020-11-22 | ···················· | ···················· | ···················· | ···················· | ···················· | ···················· |
| AB | 2262 | hCoV-19/Norway/6282/2020\|EPI_ISL_775375\|2020-11-25 | ···················· | ···················· | ···················· | ···················· | ···················· | ···················· |
| AB | 2263 | hCoV-19/Norway/6284/2020\|EPI_ISL_775376\|2020-11-22 | ···················· | ····A··············· | ···················· | ···················· | ···················· | ···················· |
| AB | 2264 | hCoV-19/Norway/6288/2020\|EPI_ISL_775377\|2020-11-24 | ···················· | ···················· | ···················· | ···················· | ···················· | ···················· |
| AB | 2265 | hCoV-19/Norway/6290/2020\|EPI_ISL_775378\|2020-10-23 | ···················· | ···················· | ···················· | ···················· | ···················· | ···················· |
| AB | 2266 | hCoV-19/Norway/6292/2020\|EPI_ISL_775379\|2020-10-23 | ···················· | ···················· | ···················· | ···················· | ···················· | ···················· |
| AB | 2267 | hCoV-19/Norway/6293/2020\|EPI_ISL_775380\|2020-10-22 | ···················· | ···················· | ···················· | ···················· | ···················· | ···················· |
| AB | 2268 | hCoV-19/Norway/6295/2020\|EPI_ISL_775381\|2020-10-23 | ···················· | ···················· | ···················· | ···················· | ···················· | ···················· |
| AB | 2269 | hCoV-19/Norway/6297/2020\|EPI_ISL_775382\|2020-10-30 | ···················· | ···················· | ···················· | ···················· | ···················· | ···················· |
| AB | 2270 | hCoV-19/Norway/6298/2020\|EPI_ISL_775383\|2020-11-29 | ···················· | ···················· | ···················· | ···················· | ···················· | ···················· |
| AB | 2271 | hCoV-19/Norway/6302/2020\|EPI_ISL_775384\|2020-11-29 | ···················· | ···················· | ···················· | ···················· | ···················· | ···················· |
| AB | 2272 | hCoV-19/Norway/6303/2020\|EPI_ISL_775385\|2020-11-30 | ···················· | ···················· | ···················· | ···················· | ···················· | ···················· |
| AB | 2273 | hCoV-19/Norway/6424/2020\|EPI_ISL_775386\|2020-12-04 | ···················· | ···················· | ···················· | ···················· | ······T············· | ···················· |
| AB | 2274 | hCoV-19/Norway/6429/2020\|EPI_ISL_775387\|2020-11-25 | ···················· | ·A·················· | ···················· | ···················· | ···················· | ···A················ |
| AB | 2275 | hCoV-19/Norway/6430/2020\|EPI_ISL_775388\|2020-11-18 | ···················· | ·A·················· | ···················· | ···················· | ···················· | ···A················ |
| AB | 2276 | hCoV-19/Norway/6432/2020\|EPI_ISL_775389\|2020-11-28 | ···················· | ·A·················· | ···················· | ···················· | ···················· | ···A················ |
| AB | 2277 | hCoV-19/Norway/6433/2020\|EPI_ISL_775390\|2020-11-25 | ···················· | ·A·················· | ···················· | ···················· | ···················· | ···A················ |
| AB | 2278 | hCoV-19/Norway/6456/2020\|EPI_ISL_775391\|2020-12-03 | ···················· | ···················· | ···················· | ···················· | ···················· | ···················· |
| AB | 2279 | hCoV-19/Norway/6457/2020\|EPI_ISL_775392\|2020-12-04 | ···················· | ···················· | ···················· | ···················· | ···················· | ···················· |
| AB | 2280 | hCoV-19/Norway/6527/2020\|EPI_ISL_775393\|2020-11-01 | ···················· | ···················· | ···················· | ···················· | ···················· | ···················· |
| AB | 2281 | hCoV-19/Norway/6572/2020\|EPI_ISL_775394\|2020-12-06 | ···················· | ···················· | ···················· | ···················· | ···················· | ···················· |
| AB | 2282 | hCoV-19/Norway/6573/2020\|EPI_ISL_775395\|2020-12-06 | ···················· | ···················· | ···················· | ···················· | ···················· | ···················· |
| AB | 2283 | hCoV-19/Norway/6574/2020\|EPI_ISL_775396\|2020-12-06 | ···················· | ···················· | ···················· | ···················· | ···················· | ···················· |
| AB | 2284 | hCoV-19/Norway/6575/2020\|EPI_ISL_775397\|2020-12-06 | ···················· | ···················· | ···················· | ···················· | ···················· | ···················· |
| AB | 2285 | hCoV-19/Norway/6593/2020\|EPI_ISL_775398\|2020-11-23 | ···················· | ···················· | ···················· | ···················· | ···················· | ···················· |
| AB | 2286 | hCoV-19/Norway/6594/2020\|EPI_ISL_775399\|2020-12-03 | ···················· | ·A·················· | ···················· | ···················· | ···················· | ···················· |
| AB | 2287 | hCoV-19/Israel/CVL-39-ngs/2020\|EPI_ISL_776630\|2020 | ···················· | ···················· | ···················· | ···················· | ···················· | ···················· |
| AB | 2288 | hCoV-19/Israel/CVL-40-ngs/2020\|EPI_ISL_776631\|2020 | ···················· | ·········A·········· | ···················· | ···················· | ···················· | ···················· |
| AB | 2289 | hCoV-19/Israel/CVL-41-ngs/2020\|EPI_ISL_776632\|2020 | ···················· | ·········A·········· | ···················· | ···················· | ···················· | ···················· |
| AB | 2290 | hCoV-19/Israel/CVL-42-ngg/2020\|EPI_ISL_776633\|2020 | ···················· | ···················· | ···················· | ···················· | ···················· | ···················· |
| AB | 2291 | hCoV-19/Israel/CVL-47-ngs/2020\|EPI_ISL_776634\|2020 | ···················· | ·A·················· | ···················· | ···················· | ···················· | ···················· |
| AB | 2292 | hCoV-19/USA/WA-UW-25752/2020\|EPI_ISL_776690\|2020-0 | ···················· | ···················· | ···················· | ···················· | ···················· | ···················· |
| AB | 2293 | hCoV-19/USA/WA-UW-25961/2020\|EPI_ISL_776691\|2020-0 | ···················· | ···················· | ···················· | ···················· | ···················· | ···················· |
| AB | 2294 | hCoV-19/USA/WA-UW-27010/2020\|EPI_ISL_776692\|2020-0 | ···················· | ···················· | ···················· | ···················· | ···················· | ···················· |
| AB | 2295 | hCoV-19/USA/WA-UW-2798/2020\|EPI_ISL_776693\|2020-04 | ···················· | ···················· | ···················· | ···················· | ···················· | ···················· |
| AB | 2296 | hCoV-19/USA/WA-UW-34166/2020\|EPI_ISL_776694\|2020-1 | ···················· | ·A·················· | ···················· | ···················· | ···················· | ···················· |
| AB | 2297 | hCoV-19/USA/WA-UW-3859/2020\|EPI_ISL_776695\|2020-03 | ···················· | ···················· | ···················· | ···················· | ···················· | ···················· |
| AB | 2298 | hCoV-19/USA/WA-UW-47248/2020\|EPI_ISL_776696\|2020-1 | ···················· | ·A·················· | ···················· | ···················· | ···················· | ···················· |
| AB | 2299 | hCoV-19/USA/WA-UW-47257/2020\|EPI_ISL_776697\|2020-1 | ···················· | ·A·················· | ···················· | ···················· | ···················· | ···················· |
| AB | 2300 | hCoV-19/USA/WA-UW-47265/2020\|EPI_ISL_776698\|2020-1 | ········T··········· | ·A·················· | ···················· | ···················· | ···················· | ···················· |
| AB | 2301 | hCoV-19/USA/WA-UW-47266/2020\|EPI_ISL_776699\|2020-1 | ···················· | ···················· | ···················· | ···················· | ···················· | ···················· |
| AB | 2302 | hCoV-19/USA/WA-UW-47272/2020\|EPI_ISL_776700\|2020-1 | ···················· | ···················· | ···················· | ···················· | ···················· | ···················· |
| AB | 2303 | hCoV-19/England/CAMC-CB9EF1/2020\|EPI_ISL_777961\|20 | ···················· | ···················· | ···················· | ···················· | ···················· | ···················· |
| AB | 2304 | hCoV-19/England/CAMC-CB7646/2020\|EPI_ISL_777962\|20 | ···················· | ···················· | ···················· | ···················· | ···················· | ···················· |
| AB | 2305 | hCoV-19/England/QEUH-C934CE/2020\|EPI_ISL_762105\|20 | ···················· | ···················· | ···················· | ···················· | ···················· | ···················· |
| AB | 2306 | hCoV-19/England/QEUH-C934DD/2020\|EPI_ISL_762106\|20 | ···················· | ···················· | ···················· | ···················· | ···················· | ···················· |
| AB | 2307 | hCoV-19/England/QEUH-C93507/2020\|EPI_ISL_762110\|20 | ···················· | ···················· | ···················· | ···················· | ···················· | ···················· |
| AB | 2308 | hCoV-19/England/CAMC-C9947D/2020\|EPI_ISL_762113\|20 | ···················· | ···················· | ···················· | ···················· | ···················· | ···················· |
| AB | 2309 | hCoV-19/England/QEUH-C92B9D/2020\|EPI_ISL_762127\|20 | ···················· | ···················· | ···················· | ···················· | ···················· | ···················· |

| | | | | | | | | |
|---|---|---|---|---|---|---|---|---|
| AB | 2310 hCoV-19/England/QEUH-C9323D/2020\|EPI_ISL_762134\|20 | ···················· | ···················· | ···················· | ···················· | ···················· | ···················· | |
| AB | 2311 hCoV-19/England/CAMC-C98BF1/2020\|EPI_ISL_762135\|20 | ···················· | ···················· | ···················· | ···················· | ···················· | ···················· | |
| AB | 2312 hCoV-19/England/QEUH-C92FCE/2020\|EPI_ISL_762145\|20 | ···················· | ···················· | ···················· | ···················· | ···················· | ···················· | |
| AB | 2313 hCoV-19/England/QEUH-C92FA0/2020\|EPI_ISL_762149\|20 | ···················· | ···················· | ···················· | ···················· | ······T············· | ···················· | |
| AB | 2314 hCoV-19/England/QEUH-C92E76/2020\|EPI_ISL_762150\|20 | ···················· | ···················· | ···················· | ···················· | ···················· | ···················· | |
| AB | 2315 hCoV-19/England/QEUH-C9365F/2020\|EPI_ISL_762154\|20 | ···················· | ···················· | ···················· | ···················· | ···················· | ···················· | |
| AB | 2316 hCoV-19/England/CAMC-C99079/2020\|EPI_ISL_762155\|20 | ···················· | ···················· | ···················· | ···················· | ···················· | ···················· | |
| AB | 2317 hCoV-19/England/QEUH-C934FB/2020\|EPI_ISL_762156\|20 | ···················· | ···················· | ···················· | ···················· | ···················· | ···················· | |
| AB | 2318 hCoV-19/England/QEUH-C92D88/2020\|EPI_ISL_762157\|20 | ···················· | ···················· | ···················· | ···················· | ···················· | ···················· | |
| AB | 2319 hCoV-19/England/QEUH-C92BD9/2020\|EPI_ISL_762162\|20 | ···················· | ···················· | ···················· | ···················· | ···················· | ···················· | |
| AB | 2320 hCoV-19/England/CAMC-C98B79/2020\|EPI_ISL_762163\|20 | ···················· | ···················· | ···················· | ···················· | ···················· | ···················· | |
| AB | 2321 hCoV-19/England/QEUH-C93D5A/2020\|EPI_ISL_762169\|20 | ···················· | ···················· | ···················· | ···················· | ···················· | ···················· | |
| AB | 2322 hCoV-19/Wales/PHWC-4A45CF/2020\|EPI_ISL_764711\|2020 | ···················· | ···················· | ···················· | ···················· | ···················· | ···················· | |
| AB | 2324 hCoV-19/Wales/PHWC-4A45FC/2020\|EPI_ISL_764713\|2020 | ···················· | ···················· | ···················· | ···················· | ···················· | ···················· | |
| AB | 2327 hCoV-19/Wales/PHWC-4A4635/2020\|EPI_ISL_764716\|2020 | ···················· | ···················· | ···················· | ···················· | ···················· | ···················· | |
| AB | 2328 hCoV-19/Wales/PHWC-4A4644/2020\|EPI_ISL_764717\|2020 | ···················· | ···················· | ···················· | ···················· | ···················· | ···················· | |
| AB | 2333 hCoV-19/Wales/PHWC-4A46F9/2020\|EPI_ISL_764724\|2020 | ···················· | ···················· | ···················· | ···················· | NNN················· | ···················· | 1 |
| AB | 2334 hCoV-19/Wales/PHWC-4A4705/2020\|EPI_ISL_764725\|2020 | ···················· | ···················· | ···················· | ···················· | ···················· | ···················· | |
| AB | 2335 hCoV-19/Wales/PHWC-4A4714/2020\|EPI_ISL_764726\|2020 | ···················· | ···················· | ···················· | ···················· | ···················· | ·········N·········N | 1 |
| AB | 2337 hCoV-19/Wales/PHWC-4A476F/2020\|EPI_ISL_764728\|2020 | ···················· | ···················· | ···················· | ···················· | ···················· | ···················· | |
| AB | 2338 hCoV-19/Wales/PHWC-4A479C/2020\|EPI_ISL_764729\|2020 | ···················· | ···················· | ···················· | ···················· | ···················· | ···················· | |
| AB | 2339 hCoV-19/Wales/PHWC-4A47BA/2020\|EPI_ISL_764730\|2020 | ···················· | ···················· | ···················· | ···················· | ···················· | ···················· | |
| AB | 2341 hCoV-19/Wales/PHWC-4A47D8/2020\|EPI_ISL_764732\|2020 | ···················· | ···················· | ···················· | ···················· | ···················· | ···················· | |
| AB | 2342 hCoV-19/Wales/PHWC-4A47F6/2020\|EPI_ISL_764733\|2020 | ···················· | ···················· | ···················· | ···················· | ····T··············· | ···················· | |
| AB | 2343 hCoV-19/Wales/PHWC-4A4802/2020\|EPI_ISL_764734\|2020 | ···················· | ···················· | ···················· | ···················· | ···················· | ···················· | |
| AB | 2344 hCoV-19/Wales/PHWC-4A4811/2020\|EPI_ISL_764735\|2020 | ···················· | ···················· | ···················· | ···················· | ···················· | ···················· | |
| AB | 2346 hCoV-19/Wales/PHWC-4A483F/2020\|EPI_ISL_764737\|2020 | ···················· | ···················· | ···················· | ···················· | ···················· | ···················· | |
| AB | 2348 hCoV-19/Wales/PHWC-4A486C/2020\|EPI_ISL_764739\|2020 | ···················· | ···················· | ···················· | ···················· | ···················· | ···················· | |
| AB | 2349 hCoV-19/Wales/PHWC-4A487B/2020\|EPI_ISL_764740\|2020 | ···················· | ···················· | ···················· | ···················· | ···················· | ···················· | |
| AB | 2350 hCoV-19/Wales/PHWC-4A488A/2020\|EPI_ISL_764741\|2020 | ···················· | ···················· | ···················· | ···················· | ···················· | ···················· | |
| AB | 2351 hCoV-19/Wales/PHWC-4A4899/2020\|EPI_ISL_764742\|2020 | ···················· | ···················· | ···················· | ···················· | ···················· | ···················· | |
| AB | 2352 hCoV-19/Wales/PHWC-4A48A8/2020\|EPI_ISL_764743\|2020 | ···················· | ···················· | ···················· | ···················· | ···················· | ···················· | |
| AB | 2353 hCoV-19/Wales/PHWC-4A48B7/2020\|EPI_ISL_764744\|2020 | ···················· | ···················· | ···················· | ···················· | ···················· | ···················· | |
| AB | 2354 hCoV-19/Wales/PHWC-4A48C6/2020\|EPI_ISL_764745\|2020 | ···················· | ···················· | ···················· | ···················· | ···················· | ···················· | |
| AB | 2357 hCoV-19/Wales/PHWC-4A492D/2020\|EPI_ISL_764748\|2020 | ···················· | ···················· | ···················· | ···················· | ···················· | ···················· | |
| AB | 2358 hCoV-19/Wales/PHWC-4A493C/2020\|EPI_ISL_764749\|2020 | ···················· | ···················· | ···················· | ···················· | ···················· | ···················· | |
| AB | 2359 hCoV-19/Wales/PHWC-4A494B/2020\|EPI_ISL_764750\|2020 | ···················· | ···················· | ···················· | ···················· | ···················· | ···················· | |
| AB | 2360 hCoV-19/Wales/PHWC-4A495A/2020\|EPI_ISL_764751\|2020 | ···················· | ···················· | ···················· | ···················· | ···················· | ···················· | |
| AB | 2361 hCoV-19/Wales/PHWC-4A4969/2020\|EPI_ISL_764752\|2020 | ···················· | ···················· | ···················· | ···················· | ···················· | ···················· | |
| AB | 2362 hCoV-19/Wales/PHWC-4A49B4/2020\|EPI_ISL_764753\|2020 | ···················· | ···················· | ···················· | ···················· | ···················· | ···················· | |
| AB | 2364 hCoV-19/Wales/PHWC-4A49E1/2020\|EPI_ISL_764755\|2020 | ···················· | ···················· | ···················· | ···················· | ···················· | ···················· | |
| AB | 2366 hCoV-19/Wales/PHWC-4A4A0C/2020\|EPI_ISL_764757\|2020 | ···················· | ···················· | ···················· | ···················· | ···················· | ···················· | |
| AB | 2367 hCoV-19/Wales/PHWC-4A4A1B/2020\|EPI_ISL_764758\|2020 | ···················· | ···················· | ···················· | ···················· | ···················· | ···················· | |
| AB | 2368 hCoV-19/Wales/PHWC-4A4A2A/2020\|EPI_ISL_764759\|2020 | ···················· | ···················· | ···················· | ···················· | ···················· | ···················· | |
| AB | 2369 hCoV-19/Wales/PHWC-4A4A39/2020\|EPI_ISL_764760\|2020 | ···················· | ···················· | ···················· | ···················· | ···················· | ···················· | |
| AB | 2370 hCoV-19/Wales/PHWC-4A4A48/2020\|EPI_ISL_764761\|2020 | ···················· | ···················· | ···················· | ···················· | ···················· | ···················· | |
| AB | 2371 hCoV-19/Wales/PHWC-4A4A66/2020\|EPI_ISL_764762\|2020 | ···················· | ···················· | ···················· | ···················· | ···················· | ···················· | |
| AB | 2372 hCoV-19/Wales/PHWC-4A4A75/2020\|EPI_ISL_764763\|2020 | ···················· | ···················· | ···················· | ···················· | ···················· | ···················· | |
| AB | 2373 hCoV-19/Wales/PHWC-4A4A84/2020\|EPI_ISL_764764\|2020 | ···················· | ···················· | ···················· | ···················· | ···················· | ···················· | |
| AB | 2374 hCoV-19/Wales/PHWC-4A4AA2/2020\|EPI_ISL_764765\|2020 | ···················· | ···················· | ···················· | ···················· | ···················· | ···················· | |
| AB | 2375 hCoV-19/Wales/PHWC-4A4AB1/2020\|EPI_ISL_764766\|2020 | ···················· | ···················· | ···················· | ···················· | ···················· | ···················· | |
| AB | 2376 hCoV-19/Wales/PHWC-4A4ADF/2020\|EPI_ISL_764767\|2020 | ···················· | ···················· | ···················· | ···················· | ···················· | ···················· | |
| AB | 2377 hCoV-19/Wales/PHWC-4A4AEE/2020\|EPI_ISL_764768\|2020 | ···················· | ···················· | ···················· | ···················· | ···················· | ···················· | |
| AB | 2379 hCoV-19/Wales/PHWC-4A4B63/2020\|EPI_ISL_764770\|2020 | ···················· | ···················· | ···················· | ···················· | ···················· | ···················· | |
| AB | 2382 hCoV-19/Wales/PHWC-4A4BCD/2020\|EPI_ISL_764773\|2020 | ···················· | ···················· | ···················· | ···················· | ···················· | ···················· | |
| AB | 2386 hCoV-19/Wales/PHWC-4A4D8B/2020\|EPI_ISL_764788\|2020 | ···················· | ···················· | ···················· | ···················· | ···················· | ···················· | |
| AB | 2388 hCoV-19/Wales/PHWC-4A4DA9/2020\|EPI_ISL_764790\|2020 | ···················· | ···················· | ···················· | ···················· | ···················· | ···················· | |
| AB | 2400 hCoV-19/Switzerland/ZH-ETHZ-420282/2020\|EPI_ISL_76 | ···················· | ···················· | ···················· | ···················· | ···················· | ···················· | |
| AB | 2402 hCoV-19/Switzerland/ZH-ETHZ-420343/2020\|EPI_ISL_76 | ···················· | ···················· | ···················· | ···················· | ··············K····· | ···················· | |
| AB | 2404 hCoV-19/Brazil/RS-00637/2020\|EPI_ISL_770585\|2020-1 | ···················· | ···················· | ···················· | ···················· | ···················· | ···················· | |
| AB | 2405 hCoV-19/Brazil/RS-00638/2020\|EPI_ISL_770586\|2020-1 | ···················· | ···················· | ···················· | ···················· | ···················· | ···················· | |
| AB | 2406 hCoV-19/Brazil/RS-00639/2020\|EPI_ISL_770587\|2020-1 | ···················· | ···················· | ···················· | ···················· | ···················· | ···················· | |
| AB | 2407 hCoV-19/Brazil/RS-00640/2020\|EPI_ISL_770588\|2020-1 | ···················· | ···················· | ···················· | ···················· | ···················· | ···················· | |
| AB | 2408 hCoV-19/Brazil/RS-00642/2020\|EPI_ISL_770589\|2020-1 | ···················· | ···················· | ···················· | ···················· | ···················· | ···················· | |
| AB | 2409 hCoV-19/Brazil/RS-00643/2020\|EPI_ISL_770590\|2020-1 | ···················· | ···················· | ···················· | ···················· | ···················· | ···················· | |
| AB | 2410 hCoV-19/Brazil/RS-00645/2020\|EPI_ISL_770591\|2020-1 | ···················· | ···················· | ···················· | ···················· | ···················· | ···················· | |
| AB | 2411 hCoV-19/Brazil/RS-00646/2020\|EPI_ISL_770592\|2020-1 | ···················· | ···················· | ···················· | ···················· | ···················· | ···················· | |
| AB | 2412 hCoV-19/Brazil/RS-00647/2020\|EPI_ISL_770593\|2020-1 | ···················· | ···················· | ···················· | ···················· | ···················· | ···················· | |
| AB | 2413 hCoV-19/Brazil/RS-00648/2020\|EPI_ISL_770594\|2020-1 | ···················· | ···················· | ···················· | ···················· | ···················· | ···················· | |
| AB | 2414 hCoV-19/Brazil/RS-00649/2020\|EPI_ISL_770595\|2020-1 | ···················· | ···················· | ···················· | ···················· | ···················· | ···················· | |
| AB | 2415 hCoV-19/Brazil/RS-00650/2020\|EPI_ISL_770596\|2020-1 | ···················· | ···················· | ···················· | ···················· | ···················· | ···················· | |
| AB | 2416 hCoV-19/Brazil/RS-00652/2020\|EPI_ISL_770597\|2020-1 | ···················· | ···················· | ···················· | ···················· | ···················· | ···················· | |
| AB | 2417 hCoV-19/Brazil/RS-00653/2020\|EPI_ISL_770598\|2020-1 | ···················· | ···················· | ···················· | ···················· | ···················· | ···················· | |
| AB | 2418 hCoV-19/Brazil/RS-00654/2020\|EPI_ISL_770599\|2020-1 | ···················· | ···················· | ···················· | ···················· | ···················· | ···················· | |
| AB | 2419 hCoV-19/Norway/6597/2020\|EPI_ISL_775400\|2020-12-02 | ···················· | ···················· | ···················· | ···················· | ···················· | ···················· | |
| AB | 2420 hCoV-19/Norway/6601/2020\|EPI_ISL_775401\|2020-12-03 | ···················· | ·A·················· | ···················· | ···················· | ···················· | ···················· | |
| AB | 2421 hCoV-19/Norway/6604/2020\|EPI_ISL_775402\|2020-12-01 | ···················· | ···················· | ···················· | ···················· | ···················· | ···················· | |
| AB | 2422 hCoV-19/Norway/6605/2020\|EPI_ISL_775403\|2020-12-07 | ···················· | ·A·················· | ···················· | ···················· | ···················· | ···················· | |
| AB | 2423 hCoV-19/Norway/6613/2020\|EPI_ISL_775404\|2020-12-04 | ···················· | ·A·················· | ···················· | ···················· | ···················· | ···················· | |
| AB | 2424 hCoV-19/Norway/6615/2020\|EPI_ISL_775405\|2020-12-04 | ···················· | ·A·················· | ···················· | ···················· | ···················· | ···················· | |

| | | | | | | | | |
|---|---|---|---|---|---|---|---|---|
| AB | 2425 | hCoV-19/Norway/6703/2020\|EPI_ISL_775406\|2020-12-07 | ···················· | ···················· | ····················· | ···················· | ···················· | ···················· |
| AB | 2426 | hCoV-19/Norway/6731/2020\|EPI_ISL_775407\|2020-12-06 | ···················· | ···················· | ····················· | ···················· | ···················· | ···················· |
| AB | 2427 | hCoV-19/Norway/6734/2020\|EPI_ISL_775408\|2020-12-06 | ···················· | ···················· | ····················· | ···················· | ···················· | ···················· |
| AB | 2428 | hCoV-19/Norway/6735/2020\|EPI_ISL_775409\|2020-12-05 | ···················· | ···················· | ····················· | ···················· | ···················· | ···················· |
| AB | 2429 | hCoV-19/Norway/6771/2020\|EPI_ISL_775410\|2020-12-10 | ···················· | ···················· | ····················· | ···················· | ···················· | ···················· |
| AB | 2430 | hCoV-19/Norway/6772/2020\|EPI_ISL_775411\|2020-12-12 | ···················· | ···················· | ····················· | ···················· | ···················· | ···················· |
| AB | 2431 | hCoV-19/Norway/6773/2020\|EPI_ISL_775412\|2020-12-11 | ···················· | ·A·················· | ····················· | ···················· | ···················· | ···A················ |
| AB | 2432 | hCoV-19/Norway/6774/2020\|EPI_ISL_775413\|2020-12-12 | ···················· | ···················· | ····················· | ···················· | ···················· | ···················· |
| AB | 2433 | hCoV-19/Norway/7092/2020\|EPI_ISL_775418\|2020-11-30 | ···················· | ···················· | ····················· | ···················· | ···················· | ···················· |
| AB | 2434 | hCoV-19/Norway/7102/2020\|EPI_ISL_775425\|2020-12-18 | ···················· | ···················· | ····················· | ···················· | ···················· | ···················· |
| AB | 2435 | hCoV-19/Norway/7103/2020\|EPI_ISL_775426\|2020-12-18 | ···················· | ···················· | ····················· | ···················· | ···················· | ···················· |
| AB | 2436 | hCoV-19/Norway/7108/2020\|EPI_ISL_775427\|2020-12-21 | ···················· | ·A·················· | ····················· | ···················· | ···················· | ···A················ |
| AB | 2437 | hCoV-19/Norway/7180/2020\|EPI_ISL_775428\|2020-12-27 | ···················· | ·A·················· | ····················· | ···················· | ···················· | ···A················ |
| AB | 2438 | hCoV-19/Norway/7181/2020\|EPI_ISL_775429\|2020-12-27 | ···················· | ·A·················· | ····················· | ···················· | ···················· | ···A················ |
| AB | 2439 | hCoV-19/Norway/7182/2020\|EPI_ISL_775430\|2020-12-21 | ···················· | ···················· | ····················· | ···················· | ······T············· | ···················· |
| AB | 2440 | hCoV-19/Norway/7183/2020\|EPI_ISL_775431\|2020-12-21 | ···················· | ···················· | ····················· | ···················· | ······T············· | ···················· |
| AB | 2441 | hCoV-19/Norway/7184/2020\|EPI_ISL_775432\|2020-12-27 | ···················· | ···················· | ····················· | ···················· | ······T············· | ···················· |
| AB | 2442 | hCoV-19/Norway/7185/2020\|EPI_ISL_775433\|2020-12-27 | ···················· | ···················· | ····················· | ···················· | ······T············· | ···················· |
| AB | 2443 | hCoV-19/Norway/7186/2020\|EPI_ISL_775434\|2020-12-22 | ···················· | ·A·················· | ····················· | ···················· | ···················· | ···A················ |
| AB | 2444 | hCoV-19/Norway/7187/2020\|EPI_ISL_775435\|2020-12-22 | ···················· | ···················· | ····················· | ···················· | ······T············· | ···················· |
| AB | 2445 | hCoV-19/Norway/7188/2020\|EPI_ISL_775436\|2020-12-27 | ···················· | ·A·················· | ····················· | ···················· | ···················· | ···A················ |
| AB | 2446 | hCoV-19/Norway/7189/2020\|EPI_ISL_775437\|2020-12-26 | ···················· | ···················· | ····················· | ···················· | ······T············· | ···················· |
| AB | 2447 | hCoV-19/Norway/7016/2020\|EPI_ISL_775438\|2020-12-17 | ···················· | ···················· | ····················· | ···················· | ···················· | ···················· |
| AB | 2448 | hCoV-19/Norway/7019/2020\|EPI_ISL_775439\|2020-12-18 | ···················· | ···················· | ····················· | ···················· | ···················· | ···················· |
| AB | 2449 | hCoV-19/Norway/4046/2020\|EPI_ISL_775440\|2020-10-06 | ···················· | ···················· | ····················· | ···················· | ···················· | ···················· |
| AB | 2450 | hCoV-19/Norway/4864/2020\|EPI_ISL_775441\|2020-10-19 | ···················· | ···················· | ····················· | ···················· | ···················· | ···················· |
| AB | 2451 | hCoV-19/Norway/6660/2020\|EPI_ISL_775442\|2020-12-11 | ···················· | ···················· | ····················· | ···················· | ···················· | ···················· |
| AB | 2452 | hCoV-19/Norway/6775/2020\|EPI_ISL_775443\|2020-12-06 | ···················· | ···················· | ····················· | ···················· | ···················· | ···················· |
| AB | 2453 | hCoV-19/Norway/6776/2020\|EPI_ISL_775444\|2020-12-03 | ···················· | ···················· | ····················· | ···················· | ···················· | ···················· |
| AB | 2454 | hCoV-19/Norway/4099/2020\|EPI_ISL_775445\|2020-10-03 | ···················· | ···················· | ····················· | ···················· | ···················· | ···················· |
| AB | 2455 | hCoV-19/Norway/4307/2020\|EPI_ISL_775446\|2020-10-06 | ···················· | ···················· | ····················· | ···················· | ···················· | ···················· |
| AB | 2456 | hCoV-19/Norway/4309/2020\|EPI_ISL_775447\|2020-10-07 | ···················· | ···················· | ····················· | ···················· | ···················· | ···················· |
| AB | 2457 | hCoV-19/Norway/4338/2020\|EPI_ISL_775448\|2020-10-12 | ···················· | ···················· | ····················· | ···················· | ···················· | ···················· |
| AB | 2458 | hCoV-19/Norway/4345/2020\|EPI_ISL_775449\|2020-10-08 | ···················· | ···················· | ····················· | ···················· | ···················· | ···················· |
| AB | 2459 | hCoV-19/Norway/4538/2020\|EPI_ISL_775450\|2020-10-14 | ···················· | ···················· | ····················· | ···················· | ···················· | ···················· |
| AB | 2460 | hCoV-19/Norway/4548/2020\|EPI_ISL_775451\|2020-10-16 | ···················· | ···················· | ····················· | ···················· | ···················· | ···················· |
| AB | 2461 | hCoV-19/Norway/5379/2020\|EPI_ISL_775452\|2020-10-27 | ···················· | ···················· | ····················· | ···················· | ···················· | ···················· |
| AB | 2462 | hCoV-19/Norway/5381/2020\|EPI_ISL_775453\|2020-10-28 | ···················· | ···················· | ····················· | ···················· | ···················· | ···················· |
| AB | 2463 | hCoV-19/Norway/5383/2020\|EPI_ISL_775454\|2020-10-27 | ···················· | ···················· | ····················· | ···················· | ···················· | ···················· |
| AB | 2464 | hCoV-19/Norway/5403/2020\|EPI_ISL_775455\|2020-11-02 | ···················· | ···················· | ····················· | ···················· | ···················· | ···················· |
| AB | 2465 | hCoV-19/Norway/5408/2020\|EPI_ISL_775456\|2020-10-27 | ···················· | ···················· | ····················· | ···················· | ···················· | ···················· |
| AB | 2466 | hCoV-19/Norway/6590/2020\|EPI_ISL_775457\|2020-11-17 | ···················· | ···················· | ····················· | ···················· | ···················· | ···················· |
| AB | 2467 | hCoV-19/Norway/7051/2020\|EPI_ISL_775458\|2020-12-17 | ···················· | ···················· | ····················· | ···················· | ···················· | ···················· |
| AB | 2468 | hCoV-19/Norway/4145/2020\|EPI_ISL_775459\|2020-10-09 | ···················· | ···················· | ····················· | ···················· | ···················· | ···················· |
| AB | 2469 | hCoV-19/Norway/5013/2020\|EPI_ISL_775460\|2020-10-30 | ···················· | ···················· | ····················· | ···················· | ···················· | ···················· |
| AB | 2470 | hCoV-19/Norway/5039/2020\|EPI_ISL_775461\|2020-10-30 | ···················· | ···················· | ····················· | ···················· | ···················· | ···················· |
| AB | 2471 | hCoV-19/Norway/6054/2020\|EPI_ISL_775462\|2020-11-27 | ···················· | ·A·················· | ····················· | ···················· | ···················· | ···A················ |
| AB | 2472 | hCoV-19/Norway/6704/2020\|EPI_ISL_775463\|2020-12-07 | ···················· | ···················· | ····················· | ···················· | ···················· | ···················· |
| AB | 2473 | hCoV-19/Norway/3896/2020\|EPI_ISL_775464\|2020-09-30 | ···················· | ···················· | ····················· | ···················· | ···················· | ···················· |
| AB | 2474 | hCoV-19/Norway/3898/2020\|EPI_ISL_775465\|2020-09-30 | ···················· | ···················· | ····················· | ···················· | ···················· | ···················· |
| AB | 2475 | hCoV-19/Norway/3900/2020\|EPI_ISL_775466\|2020-09-30 | ···················· | ···················· | ····················· | ···················· | ···················· | ···················· |
| AB | 2476 | hCoV-19/Norway/4037/2020\|EPI_ISL_775467\|2020-09-29 | ···················· | ···················· | ····················· | ···················· | ···················· | ···················· |
| AB | 2477 | hCoV-19/Norway/4070/2020\|EPI_ISL_775468\|2020-09-28 | ···················· | ···················· | ····················· | ···················· | ···················· | ···················· |
| AB | 2478 | hCoV-19/Norway/4327/2020\|EPI_ISL_775469\|2020-10-05 | ···················· | ···················· | ····················· | ···················· | ······T············· | ···················· |
| AB | 2479 | hCoV-19/Norway/4330/2020\|EPI_ISL_775470\|2020-10-05 | ···················· | ···················· | ····················· | ···················· | ···················· | ···················· |
| AB | 2480 | hCoV-19/Norway/4331/2020\|EPI_ISL_775471\|2020-10-04 | ···················· | ···················· | ····················· | ···················· | ···················· | ···················· |
| AB | 2481 | hCoV-19/Norway/4344/2020\|EPI_ISL_775472\|2020-10-09 | ···················· | ···················· | ····················· | ···················· | ······T············· | ···················· |
| AB | 2482 | hCoV-19/Norway/4348/2020\|EPI_ISL_775473\|2020-10-10 | ···················· | ···················· | ····················· | ···················· | ···················· | ···················· |
| AB | 2483 | hCoV-19/Norway/4542/2020\|EPI_ISL_775474\|2020-10-14 | ···················· | ···················· | ····················· | ···················· | ···················· | ···················· |
| AB | 2484 | hCoV-19/Norway/4589/2020\|EPI_ISL_775475\|2020-10-18 | ···················· | ···················· | ····················· | ···················· | ···················· | ···················· |
| AB | 2485 | hCoV-19/Norway/4593/2020\|EPI_ISL_775476\|2020-10-12 | ···················· | ···················· | ····················· | ···················· | ···················· | ···················· |
| AB | 2486 | hCoV-19/Norway/4600/2020\|EPI_ISL_775477\|2020-10-12 | ···················· | ···················· | ····················· | ···················· | ···················· | ···················· |
| AB | 2487 | hCoV-19/Norway/4601/2020\|EPI_ISL_775478\|2020-10-14 | ···················· | ···················· | ····················· | ···················· | ···················· | ···················· |
| AB | 2488 | hCoV-19/Norway/4678/2020\|EPI_ISL_775479\|2020-09-28 | ···················· | ···················· | ····················· | ···················· | ···················· | ···················· |
| AB | 2489 | hCoV-19/USA/WA-UW-47276/2020\|EPI_ISL_776701\|2020-1 | ···················· | ·A·················· | ····················· | ···················· | ···················· | ···················· |
| AB | 2490 | hCoV-19/USA/WA-UW-47279/2020\|EPI_ISL_776702\|2020-1 | ···················· | ···················· | ····················· | ···················· | ···················· | ···················· |
| AB | 2491 | hCoV-19/USA/WA-UW-47280/2020\|EPI_ISL_776703\|2020-1 | ···················· | ···················· | ····················· | ···················· | ···················· | ···················· |
| AB | 2492 | hCoV-19/USA/WA-UW-47284/2020\|EPI_ISL_776704\|2020-1 | ···················· | ···················· | ····················· | ···················· | ···················· | ···················· |
| AB | 2493 | hCoV-19/USA/WA-UW-47297/2020\|EPI_ISL_776705\|2020-1 | ···················· | ···················· | ····················· | ···················· | ···················· | ···················· |
| AB | 2494 | hCoV-19/USA/WA-UW-47323/2020\|EPI_ISL_776706\|2020-1 | ···················· | ···················· | ····················· | ···················· | ···················· | ···················· |
| AB | 2495 | hCoV-19/USA/WA-UW-47332/2020\|EPI_ISL_776707\|2020-1 | ···················· | ···················· | ····················· | ···················· | ···················· | ···················· |
| AB | 2496 | hCoV-19/USA/WA-UW-47337/2020\|EPI_ISL_776708\|2020-1 | ···················· | ···················· | ····················· | ···················· | ···················· | ···················· |
| AB | 2497 | hCoV-19/USA/WA-UW-47340/2020\|EPI_ISL_776709\|2020-1 | ···················· | ·A·················· | ····················· | ···················· | ···················· | ···················· |
| AB | 2498 | hCoV-19/USA/WA-UW-47361/2020\|EPI_ISL_776710\|2020-1 | ···················· | ···················· | ····················· | ···················· | ···················· | ···················· |
| AB | 2499 | hCoV-19/USA/WA-UW-47363/2020\|EPI_ISL_776712\|2020-1 | ········T··········· | ·A·················· | ····················· | ···················· | ···················· | ···················· |
| AB | 2500 | hCoV-19/USA/WA-UW-47365/2020\|EPI_ISL_776713\|2020-1 | ···················· | ···················· | ····················· | ···················· | ···················· | ···················· |
| AB | 2501 | hCoV-19/USA/WA-UW-47379/2020\|EPI_ISL_776714\|2020-1 | ···················· | ···················· | ····················· | ···················· | ···················· | ···················· |
| AB | 2502 | hCoV-19/USA/WA-UW-47383/2020\|EPI_ISL_776715\|2020-1 | ···················· | ···················· | ····················· | ···················· | ···················· | ···················· |
| AB | 2503 | hCoV-19/USA/WA-UW-47386/2020\|EPI_ISL_776716\|2020-1 | ···················· | ···················· | ····················· | ···················· | ···················· | ···················· |
| AB | 2504 | hCoV-19/USA/WA-UW-47389/2020\|EPI_ISL_776717\|2020-1 | ···················· | ···················· | ····················· | ···················· | ···················· | ···················· |

| | | | | | | | | |
|---|---|---|---|---|---|---|---|---|
| AB | 2505 | hCoV-19/USA/WA-UW-47393/2020\|EPI_ISL_776718\|2020-1 | ···················· | ···················· | ···················· | ···················· | ···················· | ···················· |
| AB | 2506 | hCoV-19/USA/WA-UW-47409/2020\|EPI_ISL_776719\|2020-1 | ···················· | ···················· | ···················· | ···················· | ···················· | ···················· |
| AB | 2507 | hCoV-19/USA/WA-UW-47412/2020\|EPI_ISL_776720\|2020-1 | ···················· | ·A·················· | ···················· | ···················· | ···················· | ···················· |
| AB | 2508 | hCoV-19/USA/WA-UW-47415/2020\|EPI_ISL_776721\|2020-1 | ···················· | ···················· | ···················· | ···················· | ···················· | ···················· |
| AB | 2509 | hCoV-19/USA/WA-UW-47416/2020\|EPI_ISL_776722\|2020-1 | ···················· | ···················· | ···················· | ···················· | ···················· | ···················· |
| AB | 2510 | hCoV-19/USA/WA-UW-47417/2020\|EPI_ISL_776723\|2020-1 | ···················· | ···················· | ···················· | ···················· | ···················· | ···················· |
| AB | 2511 | hCoV-19/USA/WA-UW-47418/2020\|EPI_ISL_776724\|2020-1 | ···················· | ·A·················· | ···················· | ···················· | ···················· | ···················· |
| AB | 2512 | hCoV-19/USA/WA-UW-47433/2020\|EPI_ISL_776725\|2020-1 | ···················· | ···················· | ···················· | ···················· | ···················· | ···················· |
| AB | 2513 | hCoV-19/USA/WA-UW-47437/2020\|EPI_ISL_776726\|2020-1 | ···················· | ···················· | ···················· | ···················· | ···················· | ···················· |
| AB | 2514 | hCoV-19/USA/WA-UW-47444/2020\|EPI_ISL_776727\|2020-1 | ···················· | ···················· | ···················· | ···················· | ···················· | ···················· |
| AB | 2515 | hCoV-19/USA/WA-UW-47450/2020\|EPI_ISL_776728\|2020-1 | ···················· | ···················· | ···················· | ···················· | ···················· | ···················· |
| AB | 2516 | hCoV-19/USA/WA-UW-47456/2020\|EPI_ISL_776729\|2020-1 | ···················· | ···················· | ···················· | ···················· | ···················· | ···················· |
| AB | 2517 | hCoV-19/USA/WA-UW-47458/2020\|EPI_ISL_776730\|2020-1 | ···················· | ·A·················· | ···················· | ···················· | ···················· | ···················· |
| AB | 2518 | hCoV-19/USA/WA-UW-49249/2020\|EPI_ISL_776731\|2020-1 | ···················· | ·A·················· | ···················· | ···················· | ···················· | ···················· |
| AB | 2519 | hCoV-19/USA/WA-UW-5488/2020\|EPI_ISL_776732\|2020-04 | ···················· | ···················· | ···················· | ···················· | ···················· | ···················· |
| AB | 2520 | hCoV-19/USA/WA-UW-5630/2020\|EPI_ISL_776733\|2020-04 | ···················· | ···················· | ···················· | ···················· | ···················· | ···················· |
| AB | 2521 | hCoV-19/USA/WA-UW-8164/2020\|EPI_ISL_776734\|2020-04 | ···················· | ···················· | ···················· | ···················· | ···················· | ···················· |
| AB | 2522 | hCoV-19/USA/WA-UW-8563/2020\|EPI_ISL_776735\|2020-05 | ···················· | ···················· | ···················· | ···················· | ···················· | ···················· |
| AB | 2523 | hCoV-19/USA/WA-UW-8613/2020\|EPI_ISL_776736\|2020-05 | ···················· | ···················· | ···················· | ···················· | ···················· | ···················· |
| AB | 2524 | hCoV-19/USA/WA-UW-8941/2020\|EPI_ISL_776737\|2020-05 | ···················· | ···················· | ···················· | ···················· | ···················· | ···················· |
| AB | 2525 | hCoV-19/USA/WA-UW-9323/2020\|EPI_ISL_776738\|2020-05 | ···················· | ···················· | ···················· | ···················· | ···················· | ···················· |
| AB | 2526 | hCoV-19/USA/WA-UW-9383/2020\|EPI_ISL_776739\|2020-05 | ···················· | ···················· | ···················· | ···················· | ···················· | ···················· |
| AB | 2527 | hCoV-19/USA/WA-UW-9948/2020\|EPI_ISL_776740\|2020-05 | ···················· | ···················· | ···················· | ···················· | ···················· | ···················· |
| AB | 2528 | hCoV-19/USA/WA-UW-47317/2020\|EPI_ISL_776741\|2020-1 | ···················· | ···················· | ···················· | ···················· | ···················· | ···················· |
| AB | 2529 | hCoV-19/USA/WA-UW-47430/2020\|EPI_ISL_776742\|2020-1 | ···················· | ···················· | ···················· | ···················· | ···················· | ···················· |
| AB | 2530 | hCoV-19/USA/WA-UW-47402/2020\|EPI_ISL_776743\|2020-1 | ···················· | ···················· | ···················· | ···················· | ···················· | ···················· |
| AB | 2531 | hCoV-19/USA/WA-UW-47422/2020\|EPI_ISL_776744\|2020-1 | ···················· | ···················· | ···················· | ···················· | ···················· | ···················· |
| AB | 2532 | hCoV-19/USA/WA-UW-8137/2020\|EPI_ISL_776749\|2020-04 | ···················· | ···················· | ···················· | ···················· | ···················· | ···················· |
| AB | 2533 | hCoV-19/Brazil/SP-821/2020\|EPI_ISL_776756\|2020-04- | ···················· | ···················· | ···················· | ···················· | ···················· | ···················· |
| AB | 2534 | hCoV-19/Brazil/SP-822/2020\|EPI_ISL_776757\|2020-04- | ···················· | ···················· | ···················· | ···················· | ···················· | ···················· |
| AB | 2535 | hCoV-19/Brazil/SP-823/2020\|EPI_ISL_776758\|2020-04- | ···················· | ···················· | ···················· | ···················· | ···················· | ···················· |
| AB | 2536 | hCoV-19/Brazil/SP-824/2020\|EPI_ISL_776759\|2020-04- | ···················· | ···················· | ···················· | ···················· | ···················· | ···················· |
| AB | 2537 | hCoV-19/Brazil/SP-825/2020\|EPI_ISL_776760\|2020-03- | ···················· | ···················· | ···················· | ···················· | ···················· | ···················· |
| AB | 2538 | hCoV-19/Brazil/SP-826/2020\|EPI_ISL_776761\|2020-03- | ···················· | ···················· | ···················· | ···················· | ···················· | ···················· |
| AB | 2539 | hCoV-19/Brazil/SP-827/2020\|EPI_ISL_776762\|2020-03- | ···················· | ···················· | ···················· | ···················· | ···················· | ···················· |
| AB | 2540 | hCoV-19/Brazil/SP-828/2020\|EPI_ISL_776763\|2020-03- | ···················· | ···················· | ···················· | ···················· | ···················· | ···················· |
| AB | 2541 | hCoV-19/Brazil/SP-776/2020\|EPI_ISL_776764\|2020-07- | ···················· | ···················· | ···················· | ···················· | ···················· | ···················· |
| AB | 2542 | hCoV-19/Brazil/SP-777/2020\|EPI_ISL_776765\|2020-07- | ···················· | ···················· | ···················· | ···················· | ···················· | ···················· |
| AB | 2543 | hCoV-19/Brazil/SP-870/2020\|EPI_ISL_776766\|2020-11- | ···················· | ···················· | ···················· | ···················· | ···················· | ···················· |
| AB | 2544 | hCoV-19/Brazil/SP-871/2020\|EPI_ISL_776767\|2020-11- | ···················· | ···················· | ···················· | ···················· | ···················· | ···················· |
| AB | 2545 | hCoV-19/Brazil/SP-872/2020\|EPI_ISL_776768\|2020-12- | ···················· | ···················· | ···················· | ···················· | ···················· | ···················· |
| AB | 2546 | hCoV-19/Brazil/SP-873/2020\|EPI_ISL_776769\|2020-11- | ···················· | ···················· | ···················· | ···················· | ···················· | ···················· |
| AB | 2548 | hCoV-19/USA/OH-ODH-SC35551/2020\|EPI_ISL_776774\|202 | ···················· | ···················· | ···················· | ···················· | ···················· | ···················· |
| AB | 2549 | hCoV-19/Israel/CVL-278-ngs/2020\|EPI_ISL_776777\|202 | ···················· | ·A·················· | ···················· | ···················· | ···················· | ···················· |
| AB | 2550 | hCoV-19/Israel/CVL-163-ngs/2020\|EPI_ISL_776778\|202 | ···················· | ···················· | ···················· | ···················· | ···················· | ···················· |
| AB | 2551 | hCoV-19/Israel/CVL-259-ngs/2020\|EPI_ISL_776779\|202 | ···················· | ···················· | ···················· | ···················· | ···················· | ···················· |
| AB | 2552 | hCoV-19/Israel/CVL-275-ngs/2020\|EPI_ISL_776791\|202 | ···················· | ···················· | ···················· | ···················· | ···················· | ···················· |
| AB | 2554 | hCoV-19/Israel/CVL-182-ngs/2020\|EPI_ISL_776793\|202 | ···················· | ···················· | ···················· | ···················· | ···················· | ···················· |
| AB | 2555 | hCoV-19/Israel/CVL-199-ngs/2020\|EPI_ISL_776794\|202 | ···················· | ···················· | ···················· | ···················· | ···················· | ···················· |
| AB | 2556 | hCoV-19/Israel/CVL-274-ngs/2020\|EPI_ISL_776795\|202 | ···················· | ···················· | ···················· | ···················· | ···················· | ···················· |
| AB | 2557 | hCoV-19/Israel/CVL-431-ngs/2020\|EPI_ISL_776796\|202 | ···················· | ···················· | ···················· | ···················· | ···················· | ···················· |
| AB | 2558 | hCoV-19/Israel/CVL-436-ngs/2020\|EPI_ISL_776797\|202 | ···················· | ···················· | ···················· | ···················· | ···················· | ···················· |
| AB | 2559 | hCoV-19/Israel/CVL-438-ngs/2020\|EPI_ISL_776798\|202 | ···················· | ···················· | ···················· | ···················· | ···················· | ···················· |
| AB | 2560 | hCoV-19/Israel/CVL-440-ngs/2020\|EPI_ISL_776799\|202 | ···················· | ···················· | ···················· | ···················· | ···················· | ···················· |
| AB | 2561 | hCoV-19/England/QEUH-C93473/2020\|EPI_ISL_762115\|20 | ···················· | ···················· | ···················· | ···················· | ···················· | ···················· |
| AB | 2562 | hCoV-19/England/QEUH-C93DB4/2020\|EPI_ISL_762120\|20 | ···················· | ···················· | ···················· | ···················· | ···················· | ···················· |
| AB | 2563 | hCoV-19/England/QEUH-C92D5B/2020\|EPI_ISL_762123\|20 | ···················· | ·A·················· | ···················· | ···················· | ···················· | ··A················· |
| AB | 2564 | hCoV-19/England/QEUH-C93D87/2020\|EPI_ISL_762171\|20 | ···················· | ···················· | ···················· | ···················· | ···················· | ···················· |
| AB | 2565 | hCoV-19/England/CAMC-C98FAA/2020\|EPI_ISL_762175\|20 | ···················· | ···················· | ···················· | ···················· | ···················· | ···················· |
| AB | 2566 | hCoV-19/England/QEUH-C93200/2020\|EPI_ISL_762182\|20 | ···················· | ···················· | ···················· | ···················· | ······T············· | ···················· |
| AB | 2567 | hCoV-19/England/CAMC-C994B9/2020\|EPI_ISL_762183\|20 | ···················· | ···················· | ···················· | ···················· | ···················· | ···················· |
| AB | 2568 | hCoV-19/England/QEUH-C92CD6/2020\|EPI_ISL_762184\|20 | ···················· | ···················· | ···················· | ···················· | ···················· | ···················· |
| AB | 2569 | hCoV-19/England/QEUH-C9316D/2020\|EPI_ISL_762188\|20 | ···················· | ···················· | ···················· | ···················· | ···················· | ···················· |
| AB | 2570 | hCoV-19/England/CAMC-C98F5F/2020\|EPI_ISL_762189\|20 | ···················· | ···················· | ···················· | ···················· | ···················· | ···················· |
| AB | 2572 | hCoV-19/Wales/PHWC-4A4C7F/2020\|EPI_ISL_764778\|2020 | ···················· | ···················· | ···················· | ···················· | ···················· | ···················· |
| AB | 2573 | hCoV-19/Wales/PHWC-4A4C8E/2020\|EPI_ISL_764779\|2020 | ···················· | ···················· | ···················· | ···················· | ···················· | ···················· |
| AB | 2574 | hCoV-19/Wales/PHWC-4A4CBB/2020\|EPI_ISL_764780\|2020 | ···················· | ···················· | ···················· | ···················· | ···················· | ···················· |
| AB | 2579 | hCoV-19/Wales/PHWC-4A4D30/2020\|EPI_ISL_764785\|2020 | ···················· | ···················· | ···················· | ···················· | ···················· | ···················· |
| AB | 2582 | hCoV-19/USA/DE-DHSS-F986750/2020\|EPI_ISL_767001\|20 | ···················· | ···················· | ···················· | ···················· | ···················· | ···················· |
| AB | 2583 | hCoV-19/Italy/APU-POLBA06/2020\|EPI_ISL_767043\|2020 | ···················· | ···················· | ···················· | ···················· | ···················· | ···················· |
| AB | 2584 | hCoV-19/USA/NM-DOH-2020641030/2020\|EPI_ISL_767051\| | ···················· | ···················· | ···················· | ···················· | ···················· | ···················· |
| AB | 2585 | hCoV-19/USA/NM-DOH-2020641432/2020\|EPI_ISL_767052\| | ···················· | ···················· | ···················· | ···················· | ···················· | ···················· |
| AB | 2586 | hCoV-19/USA/NM-DOH-2020641574/2020\|EPI_ISL_767053\| | ···················· | ···················· | ···················· | ···················· | ···················· | ···················· |
| AB | 2587 | hCoV-19/USA/NM-DOH-2020641414/2020\|EPI_ISL_767054\| | ···················· | ···················· | ···················· | ···················· | ···················· | ···················· |
| AB | 2588 | hCoV-19/USA/NM-DOH-2020640266/2020\|EPI_ISL_767055\| | ···················· | ···················· | ···················· | ···················· | ···················· | ···················· |
| AB | 2589 | hCoV-19/USA/NM-DOH-2020640985/2020\|EPI_ISL_767056\| | ···················· | ·A·················· | ···················· | ···················· | ···················· | ···················· |
| AB | 2590 | hCoV-19/USA/NM-DOH-2020641020/2020\|EPI_ISL_767057\| | ···················· | ·A·················· | ···················· | ···················· | ···················· | ···················· |
| AB | 2591 | hCoV-19/USA/NM-DOH-2020641094/2020\|EPI_ISL_767058\| | ···················· | ·A·················· | ···················· | ···················· | ···················· | ···················· |
| AB | 2592 | hCoV-19/USA/NM-DOH-2020641098/2020\|EPI_ISL_767059\| | ···················· | ···················· | ···················· | ···················· | ···················· | ···················· |
| AB | 2593 | hCoV-19/Switzerland/ZH-ETHZ-420644/2020\|EPI_ISL_76 | ···················· | ···················· | ···················· | ···················· | ···················· | ···················· |

```
AB  2594 hCoV-19/Switzerland/ZH-ETHZ-420337/2020|EPI_ISL_76  ···················· ···················· ···················· ···················· ···················· ····················
AB  2595 hCoV-19/Switzerland/ZH-ETHZ-420605/2020|EPI_ISL_76  ···················· ···················· ···················· ···················· ···················· ····················
AB  2596 hCoV-19/Singapore/1501/2020|EPI_ISL_768324|2020-12  ···················· ···················· ···················· ···················· ···················· ····················
AB  2597 hCoV-19/USA/LA-EVTL1055/2020|EPI_ISL_768325|2020-1  ···················· ·A·················· ···················· ···················· ···················· ····················
AB  2598 hCoV-19/USA/LA-EVTL1056/2020|EPI_ISL_768326|2020-1  ···················· ·A·················· ···················· ···················· ···················· ····················
AB  2599 hCoV-19/USA/LA-EVTL1057/2020|EPI_ISL_768327|2020-1  ···················· ···················· ···················· ···················· ···················· ····················
AB  2600 hCoV-19/USA/LA-EVTL1058/2020|EPI_ISL_768328|2020-1  ···················· ···················· ···················· ···················· ···················· ····················
AB  2601 hCoV-19/USA/LA-EVTL1059/2020|EPI_ISL_768329|2020-1  ···················· ···················· ···················· ···················· ···················· ····················
AB  2602 hCoV-19/USA/LA-EVTL1063/2020|EPI_ISL_768333|2020-1  ···················· ···················· ···················· ···················· ···················· ····················
AB  2603 hCoV-19/USA/LA-EVTL1080/2020|EPI_ISL_768350|2020-1  ···················· ···················· ···················· ···················· ···················· ····················
AB  2604 hCoV-19/USA/LA-EVTL1081/2020|EPI_ISL_768351|2020-1  ···················· ·A·················· ···················· ···················· ···················· ····················
AB  2605 hCoV-19/USA/LA-EVTL1082/2020|EPI_ISL_768352|2020-1  ···················· ···················· ···················· ···················· ···················· ····················
AB  2606 hCoV-19/USA/LA-EVTL1083/2020|EPI_ISL_768353|2020-1  ···················· ···················· ···················· ···················· ···················· ····················
AB  2607 hCoV-19/USA/LA-EVTL1084/2020|EPI_ISL_768354|2020-1  ···················· ···················· ···················· ···················· ···················· ····················
AB  2608 hCoV-19/USA/LA-EVTL1085/2020|EPI_ISL_768355|2020-1  ···················· ···················· ···················· ···················· ···················· ····················
AB  2609 hCoV-19/USA/LA-EVTL1086/2020|EPI_ISL_768356|2020-1  ···················· ···················· ···················· ···················· ···················· ····················
AB  2610 hCoV-19/USA/LA-EVTL1087/2020|EPI_ISL_768357|2020-1  ···················· ···················· ···················· ···················· ···················· ····················
AB  2611 hCoV-19/USA/LA-EVTL1088/2020|EPI_ISL_768358|2020-1  ···················· ···················· ···················· ···················· ···················· ····················
AB  2612 hCoV-19/USA/LA-EVTL1089/2020|EPI_ISL_768359|2020-1  ···················· ···················· ···················· ···················· ···················· ····················
AB  2613 hCoV-19/USA/LA-EVTL1090/2020|EPI_ISL_768360|2020-1  ···················· ···················· ···················· ···················· ···················· ····················
AB  2614 hCoV-19/USA/LA-EVTL1091/2020|EPI_ISL_768361|2020-1  ···················· ···················· ···················· ···················· ···················· ····················
AB  2615 hCoV-19/USA/LA-EVTL1092/2020|EPI_ISL_768362|2020-1  ···················· ···················· ···················· ···················· ···················· ····················
AB  2616 hCoV-19/USA/LA-EVTL1093/2020|EPI_ISL_768363|2020-1  ···················· ···················· ···················· ···················· ···················· ····················
AB  2617 hCoV-19/USA/LA-EVTL1095/2020|EPI_ISL_768365|2020-1  ···················· ···················· ···················· ···················· ···················· ····················
AB  2618 hCoV-19/USA/LA-EVTL1096/2020|EPI_ISL_768366|2020-1  ···················· ·A·················· ···················· ···················· ···················· ····················
AB  2619 hCoV-19/USA/LA-EVTL1097/2020|EPI_ISL_768367|2020-1  ···················· ·A·················· ···················· ···················· ···················· ····················
AB  2620 hCoV-19/USA/LA-EVTL1098/2020|EPI_ISL_768368|2020-1  ···················· ···················· ···················· ···················· ···················· ····················
AB  2621 hCoV-19/USA/LA-EVTL1099/2020|EPI_ISL_768369|2020-1  ···················· ·A·················· ···················· ···················· ···················· ····················
AB  2622 hCoV-19/USA/LA-EVTL1100/2020|EPI_ISL_768370|2020-1  ···················· ·A·················· ···················· ···················· ···················· ····················
AB  2623 hCoV-19/USA/LA-EVTL1101/2020|EPI_ISL_768371|2020-1  ···················· ···················· ···················· ···················· ···················· ····················
AB  2624 hCoV-19/USA/LA-EVTL1102/2020|EPI_ISL_768372|2020-1  ···················· ···················· ···················· ···················· ···················· ····················
AB  2625 hCoV-19/USA/LA-EVTL1103/2020|EPI_ISL_768373|2020-1  ···················· ···················· ···················· ···················· ···················· ····················
AB  2626 hCoV-19/USA/LA-EVTL1104/2020|EPI_ISL_768374|2020-1  ···················· ···················· ···················· ···················· ···················· ····················
AB  2627 hCoV-19/USA/LA-EVTL1105/2020|EPI_ISL_768375|2020-1  ···················· ·A·················· ···················· ···················· ···················· ····················
AB  2628 hCoV-19/USA/LA-EVTL1106/2020|EPI_ISL_768376|2020-1  ···················· ···················· ···················· ···················· ···················· ····················
AB  2629 hCoV-19/USA/LA-EVTL1107/2020|EPI_ISL_768377|2020-1  ···················· ···················· ···················· ···················· ···················· ····················
AB  2630 hCoV-19/USA/LA-EVTL1108/2020|EPI_ISL_768378|2020-1  ···················· ···A················ ···················· ···················· ···················· ····················
AB  2631 hCoV-19/USA/LA-EVTL1109/2020|EPI_ISL_768379|2020-1  ···················· ···················· ···················· ···················· ···················· ····················
AB  2632 hCoV-19/USA/LA-EVTL1110/2020|EPI_ISL_768380|2020-1  ···················· ···················· ···················· ···················· ···················· ····················
AB  2633 hCoV-19/USA/LA-EVTL1111/2020|EPI_ISL_768381|2020-1  ···················· ···················· ···················· ···················· ···················· ····················
AB  2634 hCoV-19/USA/LA-EVTL1112/2020|EPI_ISL_768382|2020-1  ···················· ···················· ···················· ···················· ···················· ····················
AB  2635 hCoV-19/USA/LA-EVTL1113/2020|EPI_ISL_768383|2020-1  ···················· ···················· ···················· ···················· ···················· ····················
AB  2636 hCoV-19/USA/LA-EVTL1114/2020|EPI_ISL_768384|2020-1  ···················· ·A·················· ···················· ···················· ···················· ····················
AB  2637 hCoV-19/USA/LA-EVTL1115/2020|EPI_ISL_768385|2020-1  ···················· ···················· ···················· ···················· ···················· ····················
AB  2638 hCoV-19/USA/LA-EVTL1116/2020|EPI_ISL_768386|2020-1  ···················· ···················· ···················· ···················· ···················· ····················
AB  2639 hCoV-19/USA/LA-EVTL1117/2020|EPI_ISL_768387|2020-1  ···················· ···················· ···················· ···················· ···················· ····················
AB  2640 hCoV-19/USA/LA-EVTL1118/2020|EPI_ISL_768388|2020-1  ···················· ···················· ···················· ···················· ···················· ····················
AB  2641 hCoV-19/USA/LA-EVTL1119/2020|EPI_ISL_768389|2020-1  ···················· ···················· ···················· ···················· ···················· ····················
AB  2642 hCoV-19/England/MILK-C53415/2020|EPI_ISL_769602|20  ···················· ···················· ···················· ···················· ···················· ····················
AB  2643 hCoV-19/England/MILK-C51893/2020|EPI_ISL_769603|20  ···················· ·A·················· ···················· ···················· ···················· ···A················
AB  2644 hCoV-19/England/QEUH-C4B717/2020|EPI_ISL_769607|20  ···················· ·········T·········· ···················· ···················· ···················· ····················
AB  2645 hCoV-19/England/QEUH-C4BA5A/2020|EPI_ISL_769609|20  ···················· ···················· ···················· ···················· ······T············· ····················
AB  2646 hCoV-19/England/MILK-C5017B/2020|EPI_ISL_769610|20  ···················· ···················· ···················· ···················· ···················· ····················
AB  2647 hCoV-19/England/QEUH-C4B8E7/2020|EPI_ISL_769611|20  ···················· ·········T·········· ···················· ···················· ···················· ····················
AB  2648 hCoV-19/England/QEUH-C4B850/2020|EPI_ISL_769614|20  ···················· ···················· ···················· ···················· ···················· ····················
AB  2649 hCoV-19/England/MILK-C53873/2020|EPI_ISL_769615|20  ···················· ···················· ···················· ···················· ···················· ····················
AB  2650 hCoV-19/England/QEUH-C4B920/2020|EPI_ISL_769617|20  ···················· ···················· ···················· ···················· ···················· ····················
AB  2651 hCoV-19/England/MILK-C5021E/2020|EPI_ISL_769618|20  ···················· ···················· ···················· ···················· ···················· ····················
AB  2652 hCoV-19/England/QEUH-C4B6FC/2020|EPI_ISL_769619|20  ···················· ···················· ···················· ···················· ···················· ····················
AB  2653 hCoV-19/England/MILK-C50269/2020|EPI_ISL_769624|20  ···················· ···················· ···················· ···················· ···················· ····················
AB  2654 hCoV-19/England/QEUH-C4B911/2020|EPI_ISL_769625|20  ···················· ···················· ···················· ···················· ···················· ····················
AB  2655 hCoV-19/England/MILK-C5175A/2020|EPI_ISL_769628|20  ···················· ···················· ···················· ···················· ···················· ····················
AB  2656 hCoV-19/England/MILK-C538FB/2020|EPI_ISL_769632|20  ···················· ···················· ···················· ···················· ···················· ····················
AB  2657 hCoV-19/England/MILK-C516B7/2020|EPI_ISL_769635|20  ···················· ···················· ···················· ···················· ···················· ····················
AB  2658 hCoV-19/England/QEUH-C4BB0C/2020|EPI_ISL_769641|20  ···················· ···················· ···················· ···················· ···················· ····················
AB  2659 hCoV-19/England/MILK-C502C3/2020|EPI_ISL_769642|20  ···················· ···················· ···················· ···················· ···················· ····················
AB  2660 hCoV-19/England/MILK-C51857/2020|EPI_ISL_769643|20  ···················· ···················· ···················· ···················· ······T············· ····················
AB  2661 hCoV-19/England/MILK-C51620/2020|EPI_ISL_769645|20  ···················· ···················· ···················· ···················· ···················· ····················
AB  2662 hCoV-19/England/MILK-C538A0/2020|EPI_ISL_769646|20  ···················· ···················· ···················· ···················· ···················· ····················
AB  2663 hCoV-19/England/MILK-C53882/2020|EPI_ISL_769647|20  ···················· ···················· ···················· ···················· ···················· ····················
AB  2664 hCoV-19/England/MILK-C51945/2020|EPI_ISL_769649|20  ···················· ···················· ···················· ···················· ······T············· ····················
AB  2665 hCoV-19/England/QEUH-C4B841/2020|EPI_ISL_769652|20  ···················· ···················· ···················· ···················· ···················· ····················
AB  2666 hCoV-19/England/MILK-C51839/2020|EPI_ISL_769656|20  ···················· ···················· ···················· ···················· ···················· ····················
AB  2667 hCoV-19/England/MILK-C51505/2020|EPI_ISL_769664|20  ···················· ···················· ···················· ···················· ···················· ····················
AB  2668 hCoV-19/England/MILK-C51699/2020|EPI_ISL_769681|20  ···················· ···················· ···················· ···················· ···················· ····················
AB  2669 hCoV-19/England/MILK-C53A04/2020|EPI_ISL_769682|20  ···················· ···················· ···················· ···················· ···················· ····················
AB  2670 hCoV-19/England/MILK-C4FF1B/2020|EPI_ISL_769684|20  ···················· ···················· ···················· ···················· ···················· ····················
AB  2671 hCoV-19/England/MILK-C5024B/2020|EPI_ISL_769685|20  ···················· ···················· ···················· ···················· ···················· ····················
AB  2672 hCoV-19/England/QEUH-C4B735/2020|EPI_ISL_769687|20  ···················· ···················· ···················· ···················· ···················· ····················
AB  2673 hCoV-19/England/MILK-C501B7/2020|EPI_ISL_769688|20  ···················· ···················· ···················· ···················· ···················· ····················
```

| | | | | | | | |
|---|---|---|---|---|---|---|---|
| AB | 2674 hCoV-19/England/QEUH-C4B87E/2020\|EPI_ISL_769689\|20 | ···················· | ···················· | ···················· | ···················· | ···················· | ···················· |
| AB | 2675 hCoV-19/England/QEUH-C4B5FF/2020\|EPI_ISL_769691\|20 | ···················· | ···················· | ···················· | ···················· | ···················· | ···················· |
| AB | 2676 hCoV-19/England/MILK-C5361F/2020\|EPI_ISL_769692\|20 | ···················· | ···················· | ···················· | ···················· | ···················· | ···················· |
| AB | 2677 hCoV-19/England/MILK-C4FE4B/2020\|EPI_ISL_769698\|20 | ···················· | ···················· | ···················· | ···················· | ······T············· | ···················· |
| AB | 2678 hCoV-19/Brazil/RS-00656/2020\|EPI_ISL_770600\|2020-1 | ···················· | ···················· | ···················· | ···················· | ···················· | ···················· |
| AB | 2679 hCoV-19/Brazil/RS-00657/2020\|EPI_ISL_770601\|2020-1 | ···················· | ···················· | ···················· | ···················· | ···················· | ···················· |
| AB | 2680 hCoV-19/Brazil/RS-00659/2020\|EPI_ISL_770602\|2020-1 | ···················· | ···················· | ···················· | ···················· | ···················· | ···················· |
| AB | 2681 hCoV-19/Brazil/RS-00660/2020\|EPI_ISL_770603\|2020-1 | ···················· | ···················· | ···················· | ···················· | ···················· | ···················· |
| AB | 2682 hCoV-19/Brazil/RS-00661/2020\|EPI_ISL_770604\|2020-1 | ···················· | ···················· | ···················· | ···················· | ···················· | ···················· |
| AB | 2683 hCoV-19/Brazil/RS-00662/2020\|EPI_ISL_770605\|2020-1 | ···················· | ···················· | ···················· | ···················· | ···················· | ···················· |
| AB | 2684 hCoV-19/Brazil/RS-00663/2020\|EPI_ISL_770606\|2020-1 | ···················· | ···················· | ···················· | ···················· | ···················· | ···················· |
| AB | 2685 hCoV-19/Brazil/RS-00665/2020\|EPI_ISL_770607\|2020-1 | ···················· | ···················· | ···················· | ···················· | ···················· | ···················· |
| AB | 2686 hCoV-19/Brazil/RS-00666/2020\|EPI_ISL_770608\|2020-1 | ···················· | ···················· | ···················· | ···················· | ···················· | ···················· |
| AB | 2687 hCoV-19/Brazil/RS-00667/2020\|EPI_ISL_770609\|2020-1 | ···················· | ···················· | ···················· | ···················· | ···················· | ···················· |
| AB | 2688 hCoV-19/Brazil/RS-00668/2020\|EPI_ISL_770610\|2020-1 | ···················· | ···················· | ···················· | ···················· | ···················· | ···················· |
| AB | 2689 hCoV-19/Brazil/RS-00669/2020\|EPI_ISL_770611\|2020-1 | ···················· | ···················· | ···················· | ···················· | ···················· | ···················· |
| AB | 2690 hCoV-19/Brazil/RS-00670/2020\|EPI_ISL_770612\|2020-1 | ···················· | ···················· | ···················· | ···················· | ···················· | ···················· |
| AB | 2691 hCoV-19/Brazil/RS-00672/2020\|EPI_ISL_770613\|2020-1 | ···················· | ···················· | ···················· | ···················· | ···················· | ···················· |
| AB | 2692 hCoV-19/Brazil/RS-00673/2020\|EPI_ISL_770614\|2020-1 | ···················· | ···················· | ···················· | ···················· | ···················· | ···················· |
| AB | 2693 hCoV-19/Brazil/RS-00674/2020\|EPI_ISL_770615\|2020-1 | ···················· | ···················· | ···················· | ···················· | ···················· | ···················· |
| AB | 2694 hCoV-19/Brazil/RS-00675/2020\|EPI_ISL_770616\|2020-1 | ···················· | ···················· | ···················· | ···················· | ···················· | ···················· |
| AB | 2695 hCoV-19/Brazil/RS-00676/2020\|EPI_ISL_770617\|2020-1 | ···················· | ···················· | ···················· | ···················· | ···················· | ···················· |
| AB | 2696 hCoV-19/Brazil/RS-00679/2020\|EPI_ISL_770618\|2020-1 | ···················· | ···················· | ···················· | ···················· | ···················· | ···················· |
| AB | 2697 hCoV-19/Brazil/RS-00680/2020\|EPI_ISL_770619\|2020-1 | ···················· | ···················· | ···················· | ···················· | ···················· | ···················· |
| AB | 2698 hCoV-19/Brazil/RS-00681/2020\|EPI_ISL_770620\|2020-1 | ···················· | ···················· | ···················· | ···················· | ···················· | ···················· |
| AB | 2699 hCoV-19/Brazil/RS-00683/2020\|EPI_ISL_770621\|2020-1 | ···················· | ···················· | ···················· | ···················· | ···················· | ···················· |
| AB | 2700 hCoV-19/Brazil/RS-00684/2020\|EPI_ISL_770622\|2020-1 | ···················· | ···················· | ···················· | ···················· | ···················· | ···················· |
| AB | 2701 hCoV-19/Brazil/RS-00685/2020\|EPI_ISL_770623\|2020-1 | ···················· | ···················· | ···················· | ···················· | ···················· | ···················· |
| AB | 2702 hCoV-19/Brazil/RS-00686/2020\|EPI_ISL_770624\|2020-1 | ···················· | ···················· | ···················· | ···················· | ···················· | ···················· |
| AB | 2703 hCoV-19/Brazil/RS-00689/2020\|EPI_ISL_770625\|2020-1 | ···················· | ···················· | ···················· | ···················· | ···················· | ···················· |
| AB | 2704 hCoV-19/Brazil/RS-00690/2020\|EPI_ISL_770626\|2020-1 | ···················· | ···················· | ···················· | ···················· | ···················· | ···················· |
| AB | 2705 hCoV-19/Brazil/RS-00691/2020\|EPI_ISL_770627\|2020-1 | ···················· | ···················· | ···················· | ···················· | ···················· | ···················· |
| AB | 2706 hCoV-19/Brazil/RS-00692/2020\|EPI_ISL_770628\|2020-1 | ···················· | ···················· | ···················· | ···················· | ···················· | ···A················ |
| AB | 2707 hCoV-19/Brazil/RS-00693/2020\|EPI_ISL_770629\|2020-1 | ···················· | ···················· | ···················· | ···················· | ···················· | ···················· |
| AB | 2708 hCoV-19/Brazil/RS-00694/2020\|EPI_ISL_770630\|2020-1 | ···················· | ···················· | ···················· | ···················· | ···················· | ···················· |
| AB | 2709 hCoV-19/USA/OH-ODH-SC35022/2020\|EPI_ISL_770632\|202 | ···················· | ···················· | ···················· | ···················· | ···················· | ···················· |
| AB | 2710 hCoV-19/Russia/MOW-RII-MH4088S/2020\|EPI_ISL_770670 | ···················· | ···················· | ···················· | ···················· | ···················· | ···················· |
| AB | 2711 hCoV-19/Russia/SVE-RII-MH8684S/2020\|EPI_ISL_770671 | ···················· | ···················· | ···················· | ···················· | ···················· | ···················· |
| AB | 2712 hCoV-19/Russia/SVE-RII-MH8695S/2020\|EPI_ISL_770672 | ···················· | ···················· | ···················· | ···················· | ···················· | ···················· |
| AB | 2713 hCoV-19/Russia/KYA-RII-MH10306S/2020\|EPI_ISL_77067 | ···················· | ···················· | ···················· | ···················· | ···················· | ···················· |
| AB | 2714 hCoV-19/Russia/KYA-RII-MH10309S/2020\|EPI_ISL_77067 | ···················· | ···················· | ···················· | ···················· | ···················· | ···················· |
| AB | 2715 hCoV-19/Russia/KAM-RII-MH10579S/2020\|EPI_ISL_77067 | ···················· | ···················· | ···················· | ···················· | ···················· | ···················· |
| AB | 2717 hCoV-19/Russia/KAM-RII-MH10621S/2020\|EPI_ISL_77067 | ···················· | ···················· | ···················· | ···················· | ···················· | ···················· |
| AB | 2719 hCoV-19/Russia/KAM-RII-MH10627S/2020\|EPI_ISL_77067 | ···················· | ···················· | ·········T·········· | ···················· | ···················· | ········N··········· |
| AB | 2720 hCoV-19/Russia/KAM-RII-MH10662S/2020\|EPI_ISL_77068 | ···················· | ···················· | ···················· | ···················· | ···················· | ···················· |
| AB | 2721 hCoV-19/Russia/KAM-RII-MH10671S/2020\|EPI_ISL_77068 | ···················· | ···················· | ···················· | ···················· | ···················· | ···················· |
| AB | 2722 hCoV-19/Russia/KAM-RII-MH10687S/2020\|EPI_ISL_77068 | ···················· | ····A··············· | ···················· | ···················· | ···················· | ···················· |
| AB | 2723 hCoV-19/Russia/KAM-RII-MH10701S/2020\|EPI_ISL_77068 | ···················· | ···················· | ···················· | ···················· | ···················· | ···················· |
| AB | 2724 hCoV-19/Russia/KAM-RII-MH10719S/2020\|EPI_ISL_77068 | ···················· | ···················· | ···················· | ···················· | ···················· | ···················· |
| AB | 2725 hCoV-19/Russia/KAM-RII-MH10733S/2020\|EPI_ISL_77068 | ···················· | ···················· | ···················· | ···················· | ···················· | ···················· |
| AB | 2726 hCoV-19/Russia/KAM-RII-MH10735S/2020\|EPI_ISL_77068 | ···················· | ···················· | ···················· | ···················· | ···················· | ···················· |
| AB | 2727 hCoV-19/Russia/KAM-RII-MH10739S/2020\|EPI_ISL_77068 | ···················· | ···················· | ···················· | ···················· | ···················· | ···················· |
| AB | 2728 hCoV-19/Germany/NW-HHU-457/2020\|EPI_ISL_770691\|202 | ···················· | ···················· | ···················· | ···················· | ···················· | ···················· |
| AB | 2729 hCoV-19/Germany/NW-HHU-458/2020\|EPI_ISL_770692\|202 | ···················· | ···················· | ···················· | ···················· | ···················· | ···················· |
| AB | 2730 hCoV-19/Germany/NW-HHU-459/2020\|EPI_ISL_770693\|202 | ···················· | ···················· | ···················· | ···················· | ···················· | ···················· |
| AB | 2731 hCoV-19/Germany/NW-HHU-460/2020\|EPI_ISL_770694\|202 | ···················· | ···················· | ···················· | ···················· | ···················· | ···················· |
| AB | 2732 hCoV-19/Germany/NW-HHU-461/2020\|EPI_ISL_770695\|202 | ···················· | ···················· | ···················· | ···················· | ···················· | ···················· |
| AB | 2733 hCoV-19/Germany/NW-HHU-462/2020\|EPI_ISL_770696\|202 | ···················· | ···················· | ···················· | ···················· | ···················· | ···················· |
| AB | 2734 hCoV-19/Germany/NW-HHU-463/2020\|EPI_ISL_770697\|202 | ···················· | ···················· | ···················· | ···················· | ···················· | ···················· |
| AB | 2735 hCoV-19/Germany/NW-HHU-464/2020\|EPI_ISL_770698\|202 | ···················· | ···················· | ···················· | ···················· | ···················· | ···················· |
| AB | 2736 hCoV-19/Germany/NW-HHU-465/2020\|EPI_ISL_770699\|202 | ···················· | ···················· | ···················· | ···················· | ···················· | ···················· |
| AB | 2737 hCoV-19/Norway/7041/2020\|EPI_ISL_775414\|2020-12-06 | ···················· | ··G················· | ···················· | ···················· | ···················· | ···················· |
| AB | 2738 hCoV-19/Norway/7089/2020\|EPI_ISL_775415\|2020-11-27 | ···················· | ·A·················· | ···················· | ···················· | ···················· | ···A················ |
| AB | 2739 hCoV-19/Norway/7090/2020\|EPI_ISL_775416\|2020-12-30 | ···················· | ···················· | ···················· | ···················· | ···················· | ···················· |
| AB | 2740 hCoV-19/Norway/7091/2020\|EPI_ISL_775417\|2020-11-30 | ···················· | ·A·················· | ···················· | ···················· | ···················· | ···A················ |
| AB | 2741 hCoV-19/Norway/7093/2020\|EPI_ISL_775419\|2020-11-30 | ···················· | ·A·················· | ···················· | ···················· | ···················· | ···A················ |
| AB | 2742 hCoV-19/Norway/7094/2020\|EPI_ISL_775420\|2020-11-30 | ···················· | ···················· | ···················· | ···················· | ···················· | ···················· |
| AB | 2743 hCoV-19/Norway/7096/2020\|EPI_ISL_775421\|2020-12-01 | ···················· | ···················· | ···················· | ···················· | ······T············· | ···················· |
| AB | 2744 hCoV-19/Norway/7097/2020\|EPI_ISL_775422\|2020-12-01 | ···················· | ···················· | ···················· | ···················· | ···················· | ···················· |
| AB | 2745 hCoV-19/Norway/7098/2020\|EPI_ISL_775423\|2020-12-01 | ···················· | ···················· | ···················· | ···················· | ···················· | ···················· |
| AB | 2746 hCoV-19/Norway/7100/2020\|EPI_ISL_775424\|2020-12-18 | ···················· | ···················· | ···················· | ···················· | ···················· | ···················· |
| AB | 2747 hCoV-19/Norway/4760/2020\|EPI_ISL_775480\|2020-10-21 | ···················· | ···················· | ···················· | ···················· | ···················· | ···················· |
| AB | 2748 hCoV-19/Norway/4878/2020\|EPI_ISL_775481\|2020-09-30 | ···················· | ···················· | ···················· | ···················· | ···················· | ···················· |
| AB | 2749 hCoV-19/Norway/5737/2020\|EPI_ISL_775482\|2020-10-26 | ···················· | ···················· | ···················· | ···················· | ···················· | ···················· |
| AB | 2750 hCoV-19/Norway/5747/2020\|EPI_ISL_775483\|2020-10-29 | ···················· | ···················· | ···················· | ···················· | ···················· | ···················· |
| AB | 2751 hCoV-19/Norway/5822/2020\|EPI_ISL_775484\|2020-11-19 | ···················· | ·A·················· | ···················· | ···················· | ···················· | ···A················ |
| AB | 2752 hCoV-19/Norway/5837/2020\|EPI_ISL_775485\|2020-11-13 | ···················· | ···················· | ···················· | ···················· | ···················· | ···················· |
| AB | 2753 hCoV-19/Norway/5838/2020\|EPI_ISL_775486\|2020-11-03 | ···················· | ···················· | ···················· | ···················· | ···················· | ···················· |
| AB | 2754 hCoV-19/Norway/5840/2020\|EPI_ISL_775487\|2020-11-05 | ···················· | ···················· | ···················· | ···················· | ···················· | ···················· |
| AB | 2755 hCoV-19/Norway/6035/2020\|EPI_ISL_775488\|2020-11-20 | ···················· | ···················· | ···················· | ···················· | ···················· | ···················· |

| | | | | | | | |
|---|---|---|---|---|---|---|---|
| AB | 2756 hCoV-19/Norway/6041/2020\|EPI_ISL_775489\|2020-11-19 | .................... | .................... | ..................... | .................... | .................... | .................... |
| AB | 2757 hCoV-19/Norway/6043/2020\|EPI_ISL_775490\|2020-11-17 | .................... | .................... | ..................... | .................... | .................... | .................... |
| AB | 2758 hCoV-19/Norway/6195/2020\|EPI_ISL_775491\|2020-11-27 | .................... | .................... | ..................... | .................... | .................... | .................... |
| AB | 2759 hCoV-19/Norway/6196/2020\|EPI_ISL_775492\|2020-11-30 | .................... | .................... | ..................... | .................... | .................... | .................... |
| AB | 2760 hCoV-19/Norway/6197/2020\|EPI_ISL_775493\|2020-11-21 | .................... | .................... | ..................... | .................... | .................... | .................... |
| AB | 2761 hCoV-19/Norway/6198/2020\|EPI_ISL_775494\|2020-11-22 | .................... | .................... | ..................... | .................... | .................... | .................... |
| AB | 2762 hCoV-19/Norway/6240/2020\|EPI_ISL_775495\|2020-11-24 | .................... | .................... | ..................... | .................... | .................... | .................... |
| AB | 2763 hCoV-19/Norway/6387/2020\|EPI_ISL_775496\|2020-12-03 | .................... | .A.................. | ..................... | .................... | .................... | ..A................. |
| AB | 2764 hCoV-19/Norway/6391/2020\|EPI_ISL_775497\|2020-12-03 | .................... | .A.................. | ..................... | .................... | .................... | ..A................. |
| AB | 2765 hCoV-19/Norway/6472/2020\|EPI_ISL_775498\|2020-11-18 | .................... | .................... | ..................... | .................... | ..C................. | .................... |
| AB | 2766 hCoV-19/Norway/6476/2020\|EPI_ISL_775499\|2020-11-18 | .................... | .................... | ..................... | .................... | .................... | .................... |
| AB | 2767 hCoV-19/USA/WA-UW-47372/2020\|EPI_ISL_776745\|2020-1 | .................... | .................... | ..................... | .................... | .................... | .................... |
| AB | 2768 hCoV-19/USA/WA-UW-47255/2020\|EPI_ISL_776746\|2020-1 | .................... | .A.................. | ..................... | .................... | .................... | ..A................. |
| AB | 2769 hCoV-19/USA/WA-UW-8508/2020\|EPI_ISL_776747\|2020-05 | .................... | .................... | ..................... | .................... | .................... | .................... |
| AB | 2770 hCoV-19/USA/WA-UW-8467/2020\|EPI_ISL_776748\|2020-04 | .................... | .................... | ..................... | .................... | .................... | .................... |
| AB | 2771 hCoV-19/Brazil/SP-815/2020\|EPI_ISL_776750\|2020-04- | .................... | .................... | ..................... | .................... | .................... | .................... |
| AB | 2772 hCoV-19/Brazil/SP-816/2020\|EPI_ISL_776751\|2020-03- | .................... | .................... | ..................... | .................... | .................... | .................... |
| AB | 2773 hCoV-19/Brazil/SP-817/2020\|EPI_ISL_776752\|2020-03- | .................... | .................... | ..................... | .................... | .................... | .................... |
| AB | 2774 hCoV-19/Brazil/SP-818/2020\|EPI_ISL_776753\|2020-03- | .................... | .................... | ..................... | .................... | .................... | .................... |
| AB | 2775 hCoV-19/Brazil/SP-819/2020\|EPI_ISL_776754\|2020-03- | .................... | .................... | ..................... | .................... | .................... | .................... |
| AB | 2776 hCoV-19/Brazil/SP-820/2020\|EPI_ISL_776755\|2020-04- | .................... | .................... | ..................... | .................... | .................... | .................... |
| AB | 2777 hCoV-19/USA/LA-EVTL1334/2020\|EPI_ISL_779000\|2020-1 | .................... | .................... | ..................... | .................... | .................... | .................... |
| AB | 2778 hCoV-19/USA/LA-EVTL1335/2020\|EPI_ISL_779001\|2020-1 | .................... | .................... | ..................... | .................... | .................... | .................... |
| AB | 2779 hCoV-19/USA/LA-EVTL1336/2020\|EPI_ISL_779002\|2020-1 | .................... | .................... | ..................... | .................... | .................... | .................... |
| AB | 2780 hCoV-19/USA/LA-EVTL1337/2020\|EPI_ISL_779003\|2020-1 | .................... | .................... | ..................... | .................... | .................... | .................... |
| AB | 2781 hCoV-19/USA/LA-EVTL1338/2020\|EPI_ISL_779004\|2020-1 | .................... | .................... | ..................... | .................... | .................... | .................... |
| AB | 2782 hCoV-19/USA/LA-EVTL1339/2020\|EPI_ISL_779005\|2020-1 | .................... | .................... | ..................... | .................... | .................... | .................... |
| AB | 2783 hCoV-19/USA/LA-EVTL1340/2020\|EPI_ISL_779006\|2020-1 | .................... | .................... | ..................... | .................... | .................... | .................... |
| AB | 2784 hCoV-19/USA/LA-EVTL1341/2020\|EPI_ISL_779007\|2020-1 | .................... | .................... | ..................... | .................... | .................... | .................... |
| AB | 2785 hCoV-19/USA/LA-EVTL1342/2020\|EPI_ISL_779008\|2020-1 | .................... | .................... | ..................... | .................... | .................... | .................... |
| AB | 2786 hCoV-19/USA/LA-EVTL1343/2020\|EPI_ISL_779009\|2020-1 | .................... | .................... | ..................... | .................... | .................... | .................... |
| AB | 2787 hCoV-19/USA/LA-EVTL1344/2020\|EPI_ISL_779010\|2020-1 | .................... | .................... | ..................... | .................... | .................... | .................... |
| AB | 2788 hCoV-19/USA/LA-EVTL1345/2020\|EPI_ISL_779011\|2020-1 | .................... | .................... | ..................... | .................... | .................... | .................... |
| AB | 2789 hCoV-19/USA/LA-EVTL1346/2020\|EPI_ISL_779012\|2020-1 | .................... | .................... | ..................... | .................... | .................... | .................... |
| AB | 2790 hCoV-19/USA/LA-EVTL1347/2020\|EPI_ISL_779013\|2020-1 | .................... | .................... | ..................... | .................... | .................... | .................... |
| AB | 2791 hCoV-19/USA/LA-EVTL1348/2020\|EPI_ISL_779014\|2020-1 | .................... | .................... | ..................... | .................... | .................... | .................... |
| AB | 2792 hCoV-19/USA/LA-EVTL1349/2020\|EPI_ISL_779015\|2020-1 | .................... | .................... | ..................... | .................... | .................... | .................... |
| AB | 2793 hCoV-19/USA/LA-EVTL1350/2020\|EPI_ISL_779016\|2020-1 | .................... | .................... | ..................... | .................... | .................... | .................... |
| AB | 2794 hCoV-19/USA/LA-EVTL1351/2020\|EPI_ISL_779017\|2020-1 | .................... | .................... | ..................... | .................... | .................... | .................... |
| AB | 2795 hCoV-19/USA/LA-EVTL1352/2020\|EPI_ISL_779018\|2020-1 | .................... | .................... | ..................... | .................... | .................... | .................... |
| AB | 2796 hCoV-19/USA/LA-EVTL1353/2020\|EPI_ISL_779019\|2020-1 | .................... | .................... | ..................... | .................... | .................... | .................... |
| AB | 2797 hCoV-19/USA/LA-EVTL1354/2020\|EPI_ISL_779020\|2020-1 | .................... | .................... | ..................... | .................... | .................... | .................... |
| AB | 2798 hCoV-19/USA/LA-EVTL1355/2020\|EPI_ISL_779021\|2020-1 | .................... | .................... | ..................... | .................... | .................... | .................... |
| AB | 2799 hCoV-19/USA/LA-EVTL1356/2020\|EPI_ISL_779022\|2020-1 | .................... | .................... | ..................... | .................... | .................... | .................... |
| AB | 2800 hCoV-19/USA/LA-EVTL1357/2020\|EPI_ISL_779023\|2020-1 | .................... | .................... | ..................... | .................... | .................... | .................... |
| AB | 2801 hCoV-19/USA/LA-EVTL1358/2020\|EPI_ISL_779024\|2020-1 | .................... | .................... | ..................... | .................... | .................... | .................... |
| AB | 2802 hCoV-19/USA/LA-EVTL1359/2020\|EPI_ISL_779025\|2020-1 | .................... | .................... | ..................... | .................... | .................... | .................... |
| AB | 2803 hCoV-19/USA/LA-EVTL1360/2020\|EPI_ISL_779026\|2020-1 | .................... | .................... | ..................... | .................... | .................... | .................... |
| AB | 2804 hCoV-19/USA/LA-EVTL1361/2020\|EPI_ISL_779027\|2020-1 | .................... | .................... | ..................... | .................... | .................... | .................... |
| AB | 2805 hCoV-19/USA/LA-EVTL1362/2020\|EPI_ISL_779028\|2020-1 | .................... | .................... | ..................... | .................... | .................... | .................... |
| AB | 2806 hCoV-19/USA/LA-EVTL1363/2020\|EPI_ISL_779029\|2020-1 | .................... | .................... | ..................... | .................... | .................... | .................... |
| AB | 2807 hCoV-19/USA/LA-EVTL1364/2020\|EPI_ISL_779030\|2020-1 | .................... | .................... | ..................... | .................... | .................... | .................... |
| AB | 2808 hCoV-19/USA/LA-EVTL1365/2020\|EPI_ISL_779031\|2020-1 | .................... | .................... | ..................... | .................... | .................... | .................... |
| AB | 2809 hCoV-19/USA/LA-EVTL1366/2020\|EPI_ISL_779032\|2020-1 | .................... | .................... | ..................... | .................... | .................... | .................... |
| AB | 2810 hCoV-19/USA/LA-EVTL1367/2020\|EPI_ISL_779033\|2020-1 | .................... | .................... | ..................... | .................... | .................... | .................... |
| AB | 2811 hCoV-19/USA/LA-EVTL1368/2020\|EPI_ISL_779034\|2020-1 | .................... | .................... | ..................... | .................... | .................... | .................... |
| AB | 2812 hCoV-19/USA/LA-EVTL1369/2020\|EPI_ISL_779035\|2020-1 | .................... | .................... | ..................... | .................... | .................... | .................... |
| AB | 2813 hCoV-19/USA/LA-EVTL1370/2020\|EPI_ISL_779036\|2020-1 | .................... | .................... | ..................... | .................... | .................... | .................... |
| AB | 2814 hCoV-19/USA/LA-EVTL1371/2020\|EPI_ISL_779037\|2020-1 | .................... | .................... | ..................... | .................... | .................... | .................... |
| AB | 2815 hCoV-19/USA/LA-EVTL1372/2020\|EPI_ISL_779038\|2020-1 | .................... | .................... | ..................... | .................... | .................... | .................... |
| AB | 2816 hCoV-19/USA/LA-EVTL1373/2020\|EPI_ISL_779039\|2020-1 | .................... | .................... | ..................... | .................... | .................... | .................... |
| AB | 2817 hCoV-19/England/MILK-CA9BEE/2020\|EPI_ISL_762200\|20 | .................... | .................... | ..................... | .................... | .................... | .................... |
| AB | 2822 hCoV-19/Wales/PHWC-4A4F85/2020\|EPI_ISL_764804\|2020 | .................... | .................... | ..................... | .................... | .................... | .................... |
| AB | 2823 hCoV-19/Wales/PHWC-4A4FB2/2020\|EPI_ISL_764805\|2020 | .................... | .................... | ..................... | .................... | .................... | .................... |
| AB | 2824 hCoV-19/Wales/PHWC-4A4FC1/2020\|EPI_ISL_764806\|2020 | .................... | .................... | ..................... | .................... | .................... | .................... |
| AB | 2825 hCoV-19/Wales/PHWC-4A4FD0/2020\|EPI_ISL_764807\|2020 | .................... | .................... | ..................... | .................... | .................... | .................... |
| AB | 2826 hCoV-19/Wales/PHWC-4A4FEF/2020\|EPI_ISL_764808\|2020 | .................... | .................... | ..................... | .................... | .................... | .................... |
| AB | 2827 hCoV-19/Wales/PHWC-4A4FFE/2020\|EPI_ISL_764809\|2020 | .................... | .................... | ..................... | .................... | .................... | .................... |
| AB | 2828 hCoV-19/Wales/PHWC-4A5009/2020\|EPI_ISL_764810\|2020 | .................... | .................... | ..................... | .................... | .................... | .................... |
| AB | 2830 hCoV-19/Wales/PHWC-4A5027/2020\|EPI_ISL_764812\|2020 | .................... | .................... | ..................... | .................... | .................... | .................... |
| AB | 2831 hCoV-19/Wales/PHWC-4A5036/2020\|EPI_ISL_764813\|2020 | .................... | .................... | ..................... | .................... | .................... | .................... |
| AB | 2832 hCoV-19/Wales/PHWC-4A5045/2020\|EPI_ISL_764814\|2020 | .................... | .................... | ..................... | .................... | .................... | .................... |
| AB | 2833 hCoV-19/Wales/PHWC-4A5054/2020\|EPI_ISL_764815\|2020 | .................... | .................... | ..................... | .................... | .................... | .................... |
| AB | 2834 hCoV-19/Wales/PHWC-4A5063/2020\|EPI_ISL_764816\|2020 | .................... | .................... | ..................... | .................... | .................... | .................... |
| AB | 2836 hCoV-19/Wales/PHWC-4A50DC/2020\|EPI_ISL_764820\|2020 | .................... | .................... | ..................... | .................... | .................... | .................... |
| AB | 2837 hCoV-19/Wales/PHWC-4A50FA/2020\|EPI_ISL_764821\|2020 | .................... | .................... | ..................... | .................... | .................... | .................... |
| AB | 2838 hCoV-19/Wales/PHWC-4A5106/2020\|EPI_ISL_764822\|2020 | .................... | .................... | ..................... | .................... | .................... | .................... |
| AB | 2839 hCoV-19/Wales/PHWC-4A5115/2020\|EPI_ISL_764823\|2020 | .................... | .................... | ..................... | .................... | .................... | .................... |
| AB | 2841 hCoV-19/Wales/PHWC-4A5133/2020\|EPI_ISL_764825\|2020 | .................... | .................... | ..................... | .................... | .................... | .................... |
| AB | 2843 hCoV-19/Wales/PHWC-4A5151/2020\|EPI_ISL_764827\|2020 | .................... | .................... | ..................... | .................... | .................... | .................... |

| | | | | | | | |
|---|---|---|---|---|---|---|---|
| AB | 2844 hCoV-19/Wales/PHWC-4A5160/2020\|EPI_ISL_764828\|2020 | ···················· | ···················· | ···················· | ···················· | ···················· | ···················· |
| AB | 2845 hCoV-19/Wales/PHWC-4A517F/2020\|EPI_ISL_764829\|2020 | ···················· | ···················· | ···················· | ···················· | ···················· | ···················· |
| AB | 2847 hCoV-19/Wales/PHWC-4A519D/2020\|EPI_ISL_764831\|2020 | ···················· | ···················· | ···················· | ···················· | ···················· | ···················· |
| AB | 2848 hCoV-19/Wales/PHWC-4A51AC/2020\|EPI_ISL_764832\|2020 | ···················· | ···················· | ···················· | ···················· | ···················· | ···················· |
| AB | 2850 hCoV-19/Wales/PHWC-4A51CA/2020\|EPI_ISL_764834\|2020 | ···················· | ···················· | ···················· | ···················· | ···T················ | ···················· |
| AB | 2851 hCoV-19/Wales/PHWC-4A51D9/2020\|EPI_ISL_764835\|2020 | ···················· | ···················· | ···················· | ···················· | ···················· | ···················· |
| AB | 2852 hCoV-19/Wales/PHWC-4A51E8/2020\|EPI_ISL_764836\|2020 | ···················· | ···················· | ···················· | ···················· | ···················· | ···················· |
| AB | 2853 hCoV-19/Wales/PHWC-4A51F7/2020\|EPI_ISL_764837\|2020 | ···················· | ···················· | ···················· | ···················· | ···················· | ···················· |
| AB | 2854 hCoV-19/Wales/PHWC-4A5203/2020\|EPI_ISL_764838\|2020 | ···················· | ···················· | ···················· | ···················· | ···················· | ···················· |
| AB | 2855 hCoV-19/Wales/PHWC-4A5212/2020\|EPI_ISL_764839\|2020 | ···················· | ···················· | ···················· | ···················· | ···················· | ···················· |
| AB | 2856 hCoV-19/Wales/PHWC-4A5221/2020\|EPI_ISL_764840\|2020 | ···················· | ···················· | ···················· | ···················· | ···················· | ···················· |
| AB | 2857 hCoV-19/Wales/PHWC-4A5230/2020\|EPI_ISL_764841\|2020 | ···················· | ···················· | ···················· | ···················· | ···················· | ···················· |
| AB | 2858 hCoV-19/Wales/PHWC-4A524F/2020\|EPI_ISL_764842\|2020 | ···················· | ···················· | ···················· | ···················· | ···················· | ···················· |
| AB | 2859 hCoV-19/Wales/PHWC-4A525E/2020\|EPI_ISL_764843\|2020 | ···················· | ···················· | ···················· | ···················· | ···················· | ···················· |
| AB | 2860 hCoV-19/Wales/PHWC-4A527C/2020\|EPI_ISL_764844\|2020 | ···················· | ···················· | ···················· | ···················· | ···················· | ···················· |
| AB | 2861 hCoV-19/Wales/PHWC-4A528B/2020\|EPI_ISL_764845\|2020 | ···················· | ···················· | ···················· | ···················· | ···················· | ···················· |
| AB | 2864 hCoV-19/Wales/PHWC-4A52C7/2020\|EPI_ISL_764848\|2020 | ···················· | ···················· | ···················· | ···················· | ···················· | ···················· |
| AB | 2865 hCoV-19/Wales/PHWC-4A52E5/2020\|EPI_ISL_764849\|2020 | ···················· | ···················· | ···················· | ···················· | ···················· | ···················· |
| AB | 2866 hCoV-19/Wales/PHWC-4A52F4/2020\|EPI_ISL_764850\|2020 | ···················· | ···················· | ···················· | ···················· | ···················· | ···················· |
| AB | 2867 hCoV-19/Wales/PHWC-4A532E/2020\|EPI_ISL_764851\|2020 | ···················· | ···················· | ···················· | ···················· | ···················· | ···················· |
| AB | 2869 hCoV-19/Wales/PHWC-4A534C/2020\|EPI_ISL_764853\|2020 | ···················· | ···················· | ···················· | ···················· | ···················· | ···················· |
| AB | 2870 hCoV-19/Wales/PHWC-4A535B/2020\|EPI_ISL_764854\|2020 | ···················· | ···················· | ···················· | ···················· | ···················· | ···················· |
| AB | 2871 hCoV-19/Wales/PHWC-4A536A/2020\|EPI_ISL_764855\|2020 | ···················· | ···················· | ···················· | ···················· | ···················· | ···················· |
| AB | 2874 hCoV-19/Wales/PHWC-4A5397/2020\|EPI_ISL_764858\|2020 | ···················· | ···················· | ···················· | ···················· | ···················· | ···················· |
| AB | 2875 hCoV-19/Wales/PHWC-4A53A6/2020\|EPI_ISL_764859\|2020 | ···················· | ···················· | ···················· | ···················· | ···················· | ···················· |
| AB | 2877 hCoV-19/Wales/PHWC-4A53C4/2020\|EPI_ISL_764861\|2020 | ···················· | ···················· | ···················· | ···················· | ···················· | ···················· |
| AB | 2878 hCoV-19/Wales/PHWC-4A53D3/2020\|EPI_ISL_764862\|2020 | ···················· | ···················· | ···················· | ···················· | ···················· | ···················· |
| AB | 2879 hCoV-19/Wales/PHWC-4A53F1/2020\|EPI_ISL_764863\|2020 | ···················· | ···················· | ···················· | ···················· | ···················· | ···················· |
| AB | 2881 hCoV-19/USA/DE-DHSS-FLW00410316/2020\|EPI_ISL_76700 | ···················· | ···················· | ···················· | ···················· | ···················· | ···················· |
| AB | 2882 hCoV-19/USA/DE-DHSS-F987153/2020\|EPI_ISL_767002\|20 | ···················· | ·A·················· | ···················· | ···················· | ···················· | ···················· |
| AB | 2883 hCoV-19/USA/DE-DHSS-F986920/2020\|EPI_ISL_767003\|20 | ···················· | ···················· | ···················· | ···················· | ···················· | ···················· |
| AB | 2884 hCoV-19/USA/DE-DHSS-F986809/2020\|EPI_ISL_767004\|20 | ···················· | ·A·················· | ···················· | ···················· | ···················· | ···················· |
| AB | 2885 hCoV-19/USA/DE-DHSS-F1045912/2020\|EPI_ISL_767005\|2 | ···················· | ···················· | ···················· | ···················· | ···················· | ···················· |
| AB | 2886 hCoV-19/USA/DE-DHSS-F986921/2020\|EPI_ISL_767006\|20 | ···················· | ···················· | ···················· | ···················· | ···················· | ···················· |
| AB | 2887 hCoV-19/USA/DE-DHSS-F987154/2020\|EPI_ISL_767007\|20 | ···················· | ·A·················· | ···················· | ···················· | ···················· | ···················· |
| AB | 2888 hCoV-19/USA/DE-DHSS-FLW00251915/2020\|EPI_ISL_76700 | ···················· | ·····A·············· | ···················· | ···················· | ···················· | ···················· |
| AB | 2889 hCoV-19/USA/DE-DHSS-FLW00407558/2020\|EPI_ISL_76700 | ···················· | ···················· | ···················· | ···················· | ···················· | ···················· |
| AB | 2890 hCoV-19/USA/DE-DHSS-FLW00407861/2020\|EPI_ISL_76701 | ···················· | ·A·················· | ···················· | ···················· | ···················· | ···················· |
| AB | 2891 hCoV-19/USA/DE-DHSS-FLW00403673/2020\|EPI_ISL_76701 | ···················· | ·A·················· | ···················· | ···················· | ···················· | ···················· |
| AB | 2892 hCoV-19/USA/DE-DHSS-F987155/2020\|EPI_ISL_767012\|20 | ···················· | ·A·················· | ···················· | ···················· | ···················· | ···················· |
| AB | 2893 hCoV-19/USA/NM-DOH-2020641108/2020\|EPI_ISL_767060\| | ···················· | ···················· | ···················· | ···················· | ···················· | ···················· |
| AB | 2894 hCoV-19/USA/NM-DOH-2020641111/2020\|EPI_ISL_767061\| | ···················· | ···················· | ···················· | ···················· | ···················· | ···················· |
| AB | 2895 hCoV-19/USA/NM-DOH-2020641314/2020\|EPI_ISL_767062\| | ···················· | ···················· | ···················· | ···················· | ···················· | ···················· |
| AB | 2896 hCoV-19/USA/NM-DOH-2020641408/2020\|EPI_ISL_767063\| | ···················· | ···················· | ···················· | ···················· | ···················· | ···················· |
| AB | 2897 hCoV-19/USA/NM-DOH-2020642077/2020\|EPI_ISL_767064\| | ···················· | ···················· | ···················· | ···················· | ···················· | ···················· |
| AB | 2898 hCoV-19/USA/NM-DOH-2020642119/2020\|EPI_ISL_767065\| | ···················· | ···················· | ···················· | ···················· | ···················· | ···················· |
| AB | 2899 hCoV-19/USA/NM-DOH-2020642307/2020\|EPI_ISL_767066\| | ···················· | ···················· | ···················· | ···················· | ···················· | ···················· |
| AB | 2900 hCoV-19/USA/NM-DOH-2020642445/2020\|EPI_ISL_767067\| | ···················· | ···················· | ···················· | ···················· | ···················· | ···················· |
| AB | 2901 hCoV-19/USA/NM-DOH-2020642449/2020\|EPI_ISL_767068\| | ···················· | ·A·················· | ···················· | ···················· | ···················· | ···················· |
| AB | 2902 hCoV-19/USA/NM-DOH-2020642451/2020\|EPI_ISL_767069\| | ···················· | ·A·················· | ···················· | ···················· | ···················· | ···················· |
| AB | 2903 hCoV-19/USA/NM-DOH-2020642477/2020\|EPI_ISL_767070\| | ···················· | ···················· | ···················· | ···················· | ···················· | ···················· |
| AB | 2904 hCoV-19/USA/NM-DOH-2020642478/2020\|EPI_ISL_767071\| | ···················· | ···················· | ···················· | ···················· | ···················· | ···················· |
| AB | 2905 hCoV-19/USA/NM-DOH-2020642540/2020\|EPI_ISL_767072\| | ···················· | ···················· | ···················· | ···················· | ···················· | ···················· |
| AB | 2906 hCoV-19/USA/NM-DOH-2020642973/2020\|EPI_ISL_767073\| | ···················· | ···················· | ···················· | ···················· | ···················· | ···················· |
| AB | 2907 hCoV-19/USA/NM-DOH-2020642974/2020\|EPI_ISL_767074\| | ···················· | ···················· | ···················· | ···················· | ···················· | ···················· |
| AB | 2908 hCoV-19/USA/NM-DOH-2020642977/2020\|EPI_ISL_767075\| | ···················· | ···················· | ···················· | ···················· | ···················· | ···················· |
| AB | 2909 hCoV-19/USA/NM-DOH-2020642984/2020\|EPI_ISL_767076\| | ···················· | ···················· | ···················· | ···················· | ···················· | ···················· |
| AB | 2910 hCoV-19/USA/NM-DOH-2020643020/2020\|EPI_ISL_767077\| | ···················· | ···················· | ···················· | ···················· | ···················· | ···················· |
| AB | 2911 hCoV-19/USA/NM-DOH-2020643023/2020\|EPI_ISL_767078\| | ···················· | ···················· | ···················· | ···················· | ···················· | ···················· |
| AB | 2912 hCoV-19/USA/NM-DOH-2020643040/2020\|EPI_ISL_767079\| | ···················· | ···················· | ···················· | ···················· | ···················· | ···················· |
| AB | 2913 hCoV-19/USA/NM-DOH-2020643043/2020\|EPI_ISL_767080\| | ···················· | ···················· | ···················· | ···················· | ···················· | ···················· |
| AB | 2914 hCoV-19/USA/NM-DOH-2020643045/2020\|EPI_ISL_767081\| | ···················· | ···················· | ···················· | ···················· | ···················· | ···················· |
| AB | 2915 hCoV-19/USA/NM-DOH-2020643067/2020\|EPI_ISL_767082\| | ···················· | ···················· | ···················· | ···················· | ···················· | ···················· |
| AB | 2916 hCoV-19/USA/NM-DOH-2020643068/2020\|EPI_ISL_767083\| | ···················· | ···················· | ···················· | ···················· | ···················· | ···················· |
| AB | 2917 hCoV-19/USA/NM-DOH-2020643069/2020\|EPI_ISL_767084\| | ···················· | ···················· | ···················· | ···················· | ···················· | ···················· |
| AB | 2918 hCoV-19/USA/NM-DOH-2020643093/2020\|EPI_ISL_767085\| | ···················· | ···················· | ···················· | ···················· | ···················· | ···················· |
| AB | 2919 hCoV-19/USA/NM-DOH-2020643095/2020\|EPI_ISL_767086\| | ···················· | ···················· | ···················· | ···················· | ···················· | ···················· |
| AB | 2920 hCoV-19/USA/NM-DOH-2020643097/2020\|EPI_ISL_767087\| | ···················· | ···················· | ···················· | ···················· | ···················· | ···················· |
| AB | 2921 hCoV-19/USA/NM-DOH-2020644749/2020\|EPI_ISL_767088\| | ···················· | ···················· | ···················· | ···················· | ···················· | ···················· |
| AB | 2922 hCoV-19/England/MILK-CB18CE/2020\|EPI_ISL_767090\|20 | ···················· | ·A·················· | ···················· | ···················· | ···················· | ···················· |
| AB | 2923 hCoV-19/England/ALDP-CB4209/2020\|EPI_ISL_767091\|20 | ···················· | ···················· | ···················· | ···················· | ···················· | ···················· |
| AB | 2924 hCoV-19/England/QEUH-CAD79B/2020\|EPI_ISL_767092\|20 | ···················· | ···················· | ···················· | ···················· | ···················· | ···················· |
| AB | 2925 hCoV-19/England/ALDP-CB44A9/2020\|EPI_ISL_767093\|20 | ···················· | ···················· | ···················· | ···················· | ···················· | ···················· |
| AB | 2926 hCoV-19/England/QEUH-CC7AFC/2020\|EPI_ISL_767096\|20 | ···················· | ···················· | ···················· | ···················· | ···················· | ···················· |
| AB | 2927 hCoV-19/England/MILK-CA5FF5/2020\|EPI_ISL_767098\|20 | ···················· | ···················· | ···················· | ···················· | ···················· | ··A················· |
| AB | 2928 hCoV-19/England/MILK-CA68EA/2020\|EPI_ISL_767099\|20 | ···················· | ···················· | ···················· | ···················· | ···················· | ···················· |
| AB | 2929 hCoV-19/USA/LA-EVTL1060/2020\|EPI_ISL_768330\|2020-1 | ···················· | ···················· | ···················· | ···················· | ···················· | ···················· |
| AB | 2930 hCoV-19/USA/LA-EVTL1061/2020\|EPI_ISL_768331\|2020-1 | ···················· | ···················· | ···················· | ···················· | ···················· | ···················· |
| AB | 2931 hCoV-19/USA/LA-EVTL1062/2020\|EPI_ISL_768332\|2020-1 | ···················· | ···················· | ···················· | ···················· | ···················· | ···················· |
| AB | 2932 hCoV-19/USA/LA-EVTL1064/2020\|EPI_ISL_768334\|2020-1 | ···················· | ···················· | ···················· | ···················· | ···················· | ···················· |

| | | | | | | | | |
|---|---|---|---|---|---|---|---|---|
| AB | 2933 | hCoV-19/USA/LA-EVTL1065/2020\|EPI_ISL_768335\|2020-1 | ···················· | ···················· | ···················· | ···················· | ···················· | ···················· |
| AB | 2934 | hCoV-19/USA/LA-EVTL1066/2020\|EPI_ISL_768336\|2020-1 | ···················· | ···················· | ···················· | ···················· | ···················· | ···················· |
| AB | 2935 | hCoV-19/USA/LA-EVTL1067/2020\|EPI_ISL_768337\|2020-1 | ···················· | ···················· | ···················· | ···················· | ···················· | ···················· |
| AB | 2936 | hCoV-19/USA/LA-EVTL1068/2020\|EPI_ISL_768338\|2020-1 | ···················· | ···················· | ···················· | ···················· | ···················· | ···················· |
| AB | 2937 | hCoV-19/USA/LA-EVTL1069/2020\|EPI_ISL_768339\|2020-1 | ···················· | ···················· | ···················· | ···················· | ···················· | ···················· |
| AB | 2938 | hCoV-19/USA/LA-EVTL1070/2020\|EPI_ISL_768340\|2020-1 | ···················· | ···················· | ···················· | ···················· | ···················· | ···················· |
| AB | 2939 | hCoV-19/USA/LA-EVTL1071/2020\|EPI_ISL_768341\|2020-1 | ···················· | ···················· | ···················· | ···················· | ···················· | ···················· |
| AB | 2940 | hCoV-19/USA/LA-EVTL1072/2020\|EPI_ISL_768342\|2020-1 | ···················· | ·A·················· | ···················· | ···················· | ···················· | ···················· |
| AB | 2941 | hCoV-19/USA/LA-EVTL1073/2020\|EPI_ISL_768343\|2020-1 | ···················· | ···················· | ···················· | ···················· | ···················· | ···················· |
| AB | 2942 | hCoV-19/USA/LA-EVTL1074/2020\|EPI_ISL_768344\|2020-1 | ···················· | ···················· | ···················· | ···················· | ···················· | ···················· |
| AB | 2943 | hCoV-19/USA/LA-EVTL1075/2020\|EPI_ISL_768345\|2020-1 | ···················· | ···················· | ···················· | ···················· | ···················· | ···················· |
| AB | 2944 | hCoV-19/USA/LA-EVTL1076/2020\|EPI_ISL_768346\|2020-1 | ···················· | ···················· | ···················· | ···················· | ···················· | ···················· |
| AB | 2945 | hCoV-19/USA/LA-EVTL1077/2020\|EPI_ISL_768347\|2020-1 | ···················· | ···················· | ···················· | ···················· | ···················· | ···················· |
| AB | 2946 | hCoV-19/USA/LA-EVTL1078/2020\|EPI_ISL_768348\|2020-1 | ···················· | ···················· | ···················· | ···················· | ···················· | ···················· |
| AB | 2947 | hCoV-19/USA/LA-EVTL1079/2020\|EPI_ISL_768349\|2020-1 | ···················· | ···················· | ···················· | ···················· | ···················· | ···················· |
| AB | 2948 | hCoV-19/USA/LA-EVTL1120/2020\|EPI_ISL_768390\|2020-1 | ···················· | ···················· | ···················· | ···················· | ···················· | ···················· |
| AB | 2949 | hCoV-19/USA/LA-EVTL1121/2020\|EPI_ISL_768391\|2020-1 | ···················· | ·A·················· | ···················· | ···················· | ···················· | ···················· |
| AB | 2950 | hCoV-19/USA/LA-EVTL1122/2020\|EPI_ISL_768392\|2020-1 | ···················· | ···················· | ···················· | ···················· | ···················· | ···················· |
| AB | 2951 | hCoV-19/USA/LA-EVTL1123/2020\|EPI_ISL_768393\|2020-1 | ···················· | ···················· | ···················· | ···················· | ···················· | ···················· |
| AB | 2952 | hCoV-19/USA/LA-EVTL1127/2020\|EPI_ISL_768396\|2020-1 | ···················· | ·A·················· | ···················· | ···················· | ···················· | ···················· |
| AB | 2953 | hCoV-19/USA/LA-EVTL1128/2020\|EPI_ISL_768397\|2020-1 | ···················· | ·A·················· | ···················· | ···················· | ···················· | ···················· |
| AB | 2954 | hCoV-19/USA/LA-EVTL1129/2020\|EPI_ISL_768398\|2020-1 | ···················· | ···················· | ···················· | ···················· | ···················· | ···················· |
| AB | 2955 | hCoV-19/USA/LA-EVTL1130/2020\|EPI_ISL_768399\|2020-1 | ···················· | ···················· | ···················· | ···················· | ···················· | ···················· |
| AB | 2956 | hCoV-19/England/MILK-C53AC8/2020\|EPI_ISL_769660\|20 | ···················· | ···················· | ···················· | ···················· | ···················· | ···················· |
| AB | 2957 | hCoV-19/England/QEUH-C4BB75/2020\|EPI_ISL_769661\|20 | ···················· | ···················· | ···················· | ···················· | ···················· | ···················· |
| AB | 2958 | hCoV-19/England/QEUH-C4B780/2020\|EPI_ISL_769662\|20 | ···················· | ···················· | ···················· | ···················· | ···················· | ···················· |
| AB | 2959 | hCoV-19/England/MILK-C53934/2020\|EPI_ISL_769663\|20 | ···················· | ···················· | ···················· | ···················· | ···················· | ···················· |
| AB | 2960 | hCoV-19/England/MILK-C51523/2020\|EPI_ISL_769665\|20 | ···················· | ···················· | ···················· | ···················· | ···················· | ···················· |
| AB | 2961 | hCoV-19/England/MILK-C5363D/2020\|EPI_ISL_769674\|20 | ···················· | ···················· | ···················· | ···················· | ···················· | ···················· |
| AB | 2962 | hCoV-19/England/QEUH-C4B86F/2020\|EPI_ISL_769677\|20 | ···················· | ···················· | ···················· | ···················· | ···················· | ···················· |
| AB | 2963 | hCoV-19/England/MILK-C533EB/2020\|EPI_ISL_769679\|20 | ···················· | ···················· | ···················· | ···················· | ···················· | ···················· |
| AB | 2964 | hCoV-19/Norway/6487/2020\|EPI_ISL_775500\|2020-11-09 | ···················· | ·A·················· | ···················· | ···················· | ···················· | ··A················· |
| AB | 2965 | hCoV-19/Norway/6495/2020\|EPI_ISL_775501\|2020-10-31 | ···················· | ···················· | ···················· | ···················· | ···················· | ···················· |
| AB | 2966 | hCoV-19/Norway/6515/2020\|EPI_ISL_775502\|2020-10-30 | ···················· | ·A·················· | ···················· | ···················· | ···················· | ··A················· |
| AB | 2967 | hCoV-19/Norway/6521/2020\|EPI_ISL_775503\|2020-10-30 | ···················· | ···················· | ···················· | ···················· | ···················· | ···················· |
| AB | 2968 | hCoV-19/Norway/6525/2020\|EPI_ISL_775504\|2020-11-02 | ···················· | ···················· | ···················· | ···················· | ···················· | ···················· |
| AB | 2969 | hCoV-19/Norway/6602/2020\|EPI_ISL_775505\|2020-12-08 | ···················· | ···················· | ···················· | ···················· | ···················· | ···················· |
| AB | 2970 | hCoV-19/Norway/6603/2020\|EPI_ISL_775506\|2020-12-09 | ···················· | ·A·················· | ···················· | ···················· | ···················· | ··A················· |
| AB | 2971 | hCoV-19/Norway/6606/2020\|EPI_ISL_775507\|2020-12-08 | ···················· | ·A·················· | ···················· | ···················· | ···················· | ··A················· |
| AB | 2972 | hCoV-19/Norway/6609/2020\|EPI_ISL_775508\|2020-12-08 | ········T··········· | ···················· | ···················· | ···················· | ···················· | ···················· |
| AB | 2973 | hCoV-19/Norway/6689/2020\|EPI_ISL_775509\|2020-11-11 | ···················· | ·A·················· | ···················· | ···················· | ···················· | ···················· |
| AB | 2974 | hCoV-19/Norway/6690/2020\|EPI_ISL_775510\|2020-11-15 | ···················· | ···················· | ···················· | ···················· | ···················· | ···················· |
| AB | 2975 | hCoV-19/Norway/6691/2020\|EPI_ISL_775511\|2020-11-24 | ···················· | ···················· | ···················· | ···················· | ···················· | ···················· |
| AB | 2976 | hCoV-19/Norway/6692/2020\|EPI_ISL_775512\|2020-11-11 | ···················· | ···················· | ···················· | ···················· | ···················· | ···················· |
| AB | 2977 | hCoV-19/Israel/CVL-443-ngs/2020\|EPI_ISL_776800\|202 | ···················· | ···················· | ···················· | ···················· | ···················· | ···················· |
| AB | 2978 | hCoV-19/Israel/CVL-445-ngs/2020\|EPI_ISL_776801\|202 | ···················· | ···················· | ···················· | ···················· | ···················· | ···················· |
| AB | 2979 | hCoV-19/Israel/CVL-446-ngs/2020\|EPI_ISL_776802\|202 | ···················· | ···················· | ···················· | ···················· | ···················· | ···················· |
| AB | 2980 | hCoV-19/Israel/CVL-447-ngs/2020\|EPI_ISL_776803\|202 | ···················· | ···················· | ···················· | ···················· | ···················· | ···················· |
| AB | 2981 | hCoV-19/Israel/CVL-153-ngs/2020\|EPI_ISL_776804\|202 | ···················· | ···················· | ···················· | ···················· | ···················· | ···················· |
| AB | 2982 | hCoV-19/Israel/CVL-193-ngs/2020\|EPI_ISL_776805\|202 | ···················· | ···················· | ···················· | ···················· | ···················· | ···················· |
| AB | 2983 | hCoV-19/Israel/CVL-107-ngs/2020\|EPI_ISL_776813\|202 | ···················· | ···················· | ···················· | ···················· | ···················· | ···················· |
| AB | 2984 | hCoV-19/Israel/CVL-108-ngs/2020\|EPI_ISL_776814\|202 | ···················· | ···················· | ···················· | ···················· | ···················· | ···················· |
| AB | 2985 | hCoV-19/Israel/CVL-110-ngs/2020\|EPI_ISL_776815\|202 | ···················· | ···················· | ···················· | ···················· | ···················· | ···················· |
| AB | 2986 | hCoV-19/Israel/CVL-116-ngs/2020\|EPI_ISL_776816\|202 | ···················· | ···················· | ···················· | ···················· | ···················· | ···················· |
| AB | 2987 | hCoV-19/Israel/CVL-127-ngs/2020\|EPI_ISL_776817\|202 | ···················· | ···················· | ···················· | ···················· | ···················· | ···················· |
| AB | 2988 | hCoV-19/Israel/CVL-154-ngs/2020\|EPI_ISL_776818\|202 | ···················· | ···················· | ···················· | ···················· | ···················· | ···················· |
| AB | 2989 | hCoV-19/Israel/CVL-159-ngs/2020\|EPI_ISL_776819\|202 | ···················· | ···················· | ···················· | ···················· | ···················· | ···················· |
| AB | 2990 | hCoV-19/Israel/CVL-172-ngs/2020\|EPI_ISL_776820\|202 | ···················· | ·A·················· | ···················· | ···················· | ···················· | ···················· |
| AB | 2991 | hCoV-19/Israel/CVL-179-ngs/2020\|EPI_ISL_776821\|202 | ···················· | ·A·················· | ···················· | ···················· | ···················· | ···················· |
| AB | 2992 | hCoV-19/Israel/CVL-181-ngs/2020\|EPI_ISL_776822\|202 | ···················· | ···················· | ···················· | ···················· | ···················· | ···················· |
| AB | 2993 | hCoV-19/Israel/CVL-201-ngs/2020\|EPI_ISL_776823\|202 | ···················· | ···················· | ···················· | ···················· | ···················· | ···················· |
| AB | 2994 | hCoV-19/Israel/CVL-252-ngs/2020\|EPI_ISL_776824\|202 | ···················· | ···················· | ···················· | ···················· | ···················· | ···················· |
| AB | 2995 | hCoV-19/Israel/CVL-255-ngs/2020\|EPI_ISL_776825\|202 | ···················· | ·A·················· | ···················· | ···················· | ···················· | ···················· |
| AB | 2996 | hCoV-19/Israel/CVL-261-ngs/2020\|EPI_ISL_776826\|202 | ···················· | ···················· | ···················· | ···················· | ···················· | ···················· |
| AB | 2997 | hCoV-19/Israel/CVL-262-ngs/2020\|EPI_ISL_776827\|202 | ···················· | ···················· | ···················· | ···················· | ···················· | ···················· |
| AB | 2998 | hCoV-19/Israel/CVL-265-ngs/2020\|EPI_ISL_776828\|202 | ···················· | ·A·················· | ···················· | ···················· | ···················· | ···················· |
| AB | 2999 | hCoV-19/Israel/CVL-267-ngs/2020\|EPI_ISL_776829\|202 | ···················· | ···················· | ···················· | ···················· | ···················· | ···················· |
| AB | 3000 | hCoV-19/Israel/CVL-269-ngs/2020\|EPI_ISL_776830\|202 | ···················· | ···················· | ···················· | ···················· | ···················· | ···················· |
| AB | 3001 | hCoV-19/Israel/CVL-273-ngs/2020\|EPI_ISL_776831\|202 | ···················· | ·A·················· | ···················· | ···················· | ···················· | ···················· |
| AB | 3002 | hCoV-19/Israel/CVL-276-ngs/2020\|EPI_ISL_776832\|202 | ···················· | ···················· | ···················· | ···················· | ···················· | ···················· |
| AB | 3003 | hCoV-19/Israel/CVL-282-ngs/2020\|EPI_ISL_776833\|202 | ···················· | ···················· | ···················· | ···················· | ···················· | ···················· |
| AB | 3004 | hCoV-19/Israel/CVL-285-ngs/2020\|EPI_ISL_776834\|202 | ···················· | ···················· | ···················· | ···················· | ···················· | ···················· |
| AB | 3005 | hCoV-19/Israel/CVL-293-ngs/2020\|EPI_ISL_776835\|202 | ···················· | ···················· | ···················· | ···················· | ···················· | ···················· |
| AB | 3006 | hCoV-19/Israel/CVL-109-ngs/2020\|EPI_ISL_776836\|202 | ···················· | ·A·················· | ···················· | ···················· | ···················· | ···················· |
| AB | 3007 | hCoV-19/Israel/CVL-115-ngs/2020\|EPI_ISL_776837\|202 | ···················· | ···················· | ···················· | ···················· | ···················· | ···················· |
| AB | 3008 | hCoV-19/Israel/CVL-118-ngs/2020\|EPI_ISL_776838\|202 | ···················· | ···················· | ···················· | ···················· | ······T············· | ···················· |
| AB | 3009 | hCoV-19/Israel/CVL-124-ngs/2020\|EPI_ISL_776839\|202 | ···················· | ···················· | ···················· | ···················· | ···················· | ···················· |
| AB | 3010 | hCoV-19/Israel/CVL-126-ngs/2020\|EPI_ISL_776840\|202 | ···················· | ···················· | ···················· | ···················· | ···················· | ···················· |
| AB | 3011 | hCoV-19/Israel/CVL-128-ngs/2020\|EPI_ISL_776841\|202 | ···················· | ···················· | ···················· | ···················· | ···················· | ···················· |
| AB | 3012 | hCoV-19/Israel/CVL-131-ngs/2020\|EPI_ISL_776842\|202 | ···················· | ·A·················· | ···················· | ···················· | ···················· | ···················· |

| | | | | | | | |
|---|---|---|---|---|---|---|---|
| AB | 3013 hCoV-19/Israel/CVL-143-ngs/2020\|EPI_ISL_776843\|202 | ···················· | ···················· | ···················· | ···················· | ···················· | ···················· |
| AB | 3014 hCoV-19/USA/LA-EVTL1374/2020\|EPI_ISL_779040\|2020-1 | ···················· | ···················· | ···················· | ···················· | ···················· | ···················· |
| AB | 3015 hCoV-19/USA/LA-EVTL1375/2020\|EPI_ISL_779041\|2020-1 | ···················· | ···················· | ···················· | ···················· | ···················· | ···················· |
| AB | 3016 hCoV-19/USA/LA-EVTL1376/2020\|EPI_ISL_779042\|2020-1 | ···················· | ···················· | ···················· | ···················· | ···················· | ···················· |
| AB | 3017 hCoV-19/USA/LA-EVTL1377/2020\|EPI_ISL_779043\|2020-1 | ···················· | ···················· | ···················· | ···················· | ···················· | ···················· |
| AB | 3018 hCoV-19/USA/LA-EVTL1378/2020\|EPI_ISL_779044\|2020-1 | ···················· | ···················· | ···················· | ···················· | ···················· | ···················· |
| AB | 3019 hCoV-19/USA/LA-EVTL1379/2020\|EPI_ISL_779045\|2020-1 | ···················· | ···················· | ···················· | ···················· | ···················· | ···················· |
| AB | 3020 hCoV-19/USA/LA-EVTL1380/2020\|EPI_ISL_779046\|2020-1 | ···················· | ···················· | ···················· | ···················· | ···················· | ···················· |
| AB | 3021 hCoV-19/USA/LA-EVTL1381/2020\|EPI_ISL_779047\|2020-1 | ···················· | ···················· | ···················· | ···················· | ···················· | ···················· |
| AB | 3022 hCoV-19/USA/LA-EVTL1382/2020\|EPI_ISL_779048\|2020-1 | ···················· | ···················· | ···················· | ···················· | ···················· | ···················· |
| AB | 3023 hCoV-19/USA/LA-EVTL1383/2020\|EPI_ISL_779049\|2020-1 | ···················· | ···················· | ···················· | ···················· | ···················· | ···················· |
| AB | 3024 hCoV-19/USA/LA-EVTL1384/2020\|EPI_ISL_779050\|2020-1 | ···················· | ···················· | ···················· | ···················· | ···················· | ···················· |
| AB | 3025 hCoV-19/USA/LA-EVTL1385/2020\|EPI_ISL_779051\|2020-1 | ···················· | ···················· | ···················· | ···················· | ···················· | ···················· |
| AB | 3026 hCoV-19/USA/LA-EVTL1386/2020\|EPI_ISL_779052\|2020-1 | ···················· | ···················· | ···················· | ···················· | ···················· | ···················· |
| AB | 3027 hCoV-19/USA/LA-EVTL1387/2020\|EPI_ISL_779053\|2020-1 | ···················· | ···················· | ···················· | ···················· | ···················· | ···················· |
| AB | 3028 hCoV-19/USA/LA-EVTL1388/2020\|EPI_ISL_779054\|2020-1 | ···················· | ···················· | ···················· | ···················· | ···················· | ···················· |
| AB | 3029 hCoV-19/USA/LA-EVTL1389/2020\|EPI_ISL_779055\|2020-1 | ···················· | ···················· | ···················· | ···················· | ···················· | ···················· |
| AB | 3030 hCoV-19/USA/LA-EVTL1390/2020\|EPI_ISL_779056\|2020-1 | ···················· | ···················· | ···················· | ···················· | ···················· | ···················· |
| AB | 3031 hCoV-19/USA/LA-EVTL1391/2020\|EPI_ISL_779057\|2020-1 | ···················· | ···················· | ···················· | ···················· | ···················· | ···················· |
| AB | 3032 hCoV-19/USA/LA-EVTL1392/2020\|EPI_ISL_779058\|2020-1 | ···················· | ···················· | ···················· | ···················· | ···················· | ···················· |
| AB | 3033 hCoV-19/USA/LA-EVTL1393/2020\|EPI_ISL_779059\|2020-1 | ···················· | ···················· | ···················· | ···················· | ···················· | ···················· |
| AB | 3034 hCoV-19/USA/LA-EVTL1394/2020\|EPI_ISL_779060\|2020-1 | ···················· | ···················· | ···················· | ···················· | ···················· | ···················· |
| AB | 3035 hCoV-19/USA/LA-EVTL1395/2020\|EPI_ISL_779061\|2020-1 | ···················· | ···················· | ···················· | ···················· | ···················· | ···················· |
| AB | 3036 hCoV-19/USA/LA-EVTL1396/2020\|EPI_ISL_779062\|2020-1 | ···················· | ···················· | ···················· | ···················· | ···················· | ···················· |
| AB | 3037 hCoV-19/USA/LA-EVTL1397/2020\|EPI_ISL_779063\|2020-1 | ···················· | ···················· | ···················· | ···················· | ···················· | ···················· |
| AB | 3038 hCoV-19/USA/LA-EVTL1398/2020\|EPI_ISL_779064\|2020-1 | ···················· | ···················· | ···················· | ···················· | ···················· | ···················· |
| AB | 3039 hCoV-19/USA/LA-EVTL1399/2020\|EPI_ISL_779065\|2020-1 | ···················· | ···················· | ···················· | ···················· | ···················· | ···················· |
| AB | 3040 hCoV-19/USA/LA-EVTL1400/2020\|EPI_ISL_779066\|2020-1 | ···················· | ···················· | ···················· | ···················· | ···················· | ···················· |
| AB | 3041 hCoV-19/USA/LA-EVTL1401/2020\|EPI_ISL_779067\|2020-1 | ···················· | ···················· | ···················· | ···················· | ···················· | ···················· |
| AB | 3042 hCoV-19/USA/LA-EVTL1402/2020\|EPI_ISL_779068\|2020-1 | ···················· | ···················· | ···················· | ···················· | ···················· | ···················· |
| AB | 3043 hCoV-19/USA/LA-EVTL1404/2020\|EPI_ISL_779070\|2020-1 | ···················· | ···················· | ···················· | ···················· | ···················· | ···················· |
| AB | 3044 hCoV-19/USA/LA-EVTL1405/2020\|EPI_ISL_779071\|2020-1 | ···················· | ···················· | ···················· | ···················· | ···················· | ···················· |
| AB | 3045 hCoV-19/USA/LA-EVTL1406/2020\|EPI_ISL_779072\|2020-1 | ···················· | ···················· | ···················· | ···················· | ···················· | ···················· |
| AB | 3046 hCoV-19/USA/LA-EVTL1407/2020\|EPI_ISL_779073\|2020-1 | ···················· | ···················· | ···················· | ···················· | ···················· | ···················· |
| AB | 3047 hCoV-19/USA/LA-EVTL1408/2020\|EPI_ISL_779074\|2020-1 | ···················· | ···················· | ···················· | ···················· | ···················· | ···················· |
| AB | 3048 hCoV-19/USA/LA-EVTL1409/2020\|EPI_ISL_779075\|2020-1 | ···················· | ···················· | ···················· | ···················· | ···················· | ···················· |
| AB | 3049 hCoV-19/USA/LA-EVTL1410/2020\|EPI_ISL_779076\|2020-1 | ···················· | ···················· | ···················· | ···················· | ···················· | ···················· |
| AB | 3050 hCoV-19/USA/LA-EVTL1411/2020\|EPI_ISL_779077\|2020-1 | ···················· | ···················· | ···················· | ···················· | ···················· | ···················· |
| AB | 3051 hCoV-19/USA/LA-EVTL1412/2020\|EPI_ISL_779078\|2020-1 | ···················· | ···················· | ···················· | ···················· | ···················· | ···················· |
| AB | 3052 hCoV-19/USA/LA-EVTL1413/2020\|EPI_ISL_779079\|2020-1 | ···················· | ···················· | ···················· | ···················· | ···················· | ···················· |
| AB | 3053 hCoV-19/USA/LA-EVTL1414/2020\|EPI_ISL_779080\|2020-1 | ···················· | ···················· | ···················· | ···················· | ···················· | ···················· |
| AB | 3054 hCoV-19/USA/LA-EVTL1415/2020\|EPI_ISL_779081\|2020-1 | ···················· | ···················· | ···················· | ···················· | ···················· | ···················· |
| AB | 3055 hCoV-19/USA/LA-EVTL1416/2020\|EPI_ISL_779082\|2020-1 | ···················· | ···················· | ···················· | ···················· | ···················· | ···················· |
| AB | 3056 hCoV-19/USA/LA-EVTL1417/2020\|EPI_ISL_779083\|2020-1 | ···················· | ···················· | ···················· | ···················· | ···················· | ···················· |
| AB | 3057 hCoV-19/USA/LA-EVTL1418/2020\|EPI_ISL_779084\|2020-1 | ···················· | ·A·················· | ···················· | ···················· | ···················· | ···················· |
| AB | 3058 hCoV-19/USA/LA-EVTL1419/2020\|EPI_ISL_779085\|2020-1 | ···················· | ·A·················· | ···················· | ···················· | ···················· | ···················· |
| AB | 3059 hCoV-19/USA/LA-EVTL1420/2020\|EPI_ISL_779086\|2020-1 | ···················· | ·A·················· | ···················· | ···················· | ···················· | ···················· |
| AB | 3060 hCoV-19/USA/LA-EVTL1421/2020\|EPI_ISL_779087\|2020-1 | ···················· | ·A·················· | ···················· | ···················· | ···················· | ···················· |
| AB | 3061 hCoV-19/USA/LA-EVTL1422/2020\|EPI_ISL_779088\|2020-1 | ···················· | ·A·················· | ···················· | ···················· | ···················· | ···················· |
| AB | 3062 hCoV-19/USA/LA-EVTL1423/2020\|EPI_ISL_779089\|2020-1 | ···················· | ·A·················· | ···················· | ···················· | ···················· | ···················· |
| AB | 3063 hCoV-19/USA/LA-EVTL1424/2020\|EPI_ISL_779090\|2020-1 | ···················· | ·A·················· | ···················· | ···················· | ···················· | ···················· |
| AB | 3064 hCoV-19/USA/LA-EVTL1425/2020\|EPI_ISL_779091\|2020-1 | ···················· | ·A·················· | ···················· | ···················· | ···················· | ···················· |
| AB | 3065 hCoV-19/USA/LA-EVTL1426/2020\|EPI_ISL_779092\|2020-1 | ···················· | ·A·················· | ···················· | ···················· | ···················· | ···················· |
| AB | 3066 hCoV-19/USA/LA-EVTL1427/2020\|EPI_ISL_779093\|2020-1 | ···················· | ···················· | ···················· | ···················· | ···················· | ···················· |
| AB | 3067 hCoV-19/USA/LA-EVTL1428/2020\|EPI_ISL_779094\|2020-1 | ···················· | ···················· | ···················· | ···················· | ···················· | ···················· |
| AB | 3068 hCoV-19/USA/LA-EVTL1429/2020\|EPI_ISL_779095\|2020-1 | ···················· | ···················· | ···················· | ···················· | ···················· | ···················· |
| AB | 3069 hCoV-19/USA/LA-EVTL1430/2020\|EPI_ISL_779096\|2020-1 | ···················· | ···················· | ···················· | ···················· | ···················· | ···················· |
| AB | 3070 hCoV-19/USA/LA-EVTL1431/2020\|EPI_ISL_779097\|2020-1 | ···················· | ···················· | ···················· | ···················· | ···················· | ···················· |
| AB | 3071 hCoV-19/USA/LA-EVTL1432/2020\|EPI_ISL_779098\|2020-1 | ···················· | ···················· | ···················· | ···················· | ···················· | ···················· |
| AB | 3072 hCoV-19/USA/LA-EVTL1433/2020\|EPI_ISL_779099\|2020-1 | ···················· | ···················· | ···················· | ···················· | ···················· | ···················· |
| AB | 3073 hCoV-19/England/MILK-CA95FF/2020\|EPI_ISL_762214\|20 | ···················· | ···················· | ···················· | ···················· | ···················· | ···················· |
| AB | 3074 hCoV-19/England/MILK-CA9D24/2020\|EPI_ISL_762216\|20 | ···················· | ···················· | ···················· | ···················· | ···················· | ···················· |
| AB | 3075 hCoV-19/England/MILK-CA9A2D/2020\|EPI_ISL_762217\|20 | ···················· | ···················· | ···················· | ···················· | ···················· | ···················· |
| AB | 3076 hCoV-19/England/MILK-CA9D33/2020\|EPI_ISL_762225\|20 | ···················· | ···················· | ···················· | ···················· | ···················· | ···················· |
| AB | 3077 hCoV-19/England/MILK-CA9C90/2020\|EPI_ISL_762228\|20 | ···················· | ···················· | ···················· | ···················· | ······T············· | ···················· |
| AB | 3078 hCoV-19/England/MILK-CA98BA/2020\|EPI_ISL_762233\|20 | ···················· | ···················· | ···················· | ···················· | ···················· | ···················· |
| AB | 3079 hCoV-19/England/MILK-CA979F/2020\|EPI_ISL_762236\|20 | ···················· | ···················· | ···················· | ···················· | ···················· | ···················· |
| AB | 3080 hCoV-19/England/MILK-CA4CA4/2020\|EPI_ISL_762241\|20 | ···················· | ···················· | ···················· | ···················· | ···················· | ···················· |
| AB | 3081 hCoV-19/England/MILK-CA97F9/2020\|EPI_ISL_762243\|20 | ···················· | ···················· | ···················· | ···················· | ···················· | ···················· |
| AB | 3082 hCoV-19/England/MILK-CA4B6B/2020\|EPI_ISL_762244\|20 | ···················· | ···················· | ···················· | ···················· | ···················· | ···················· |
| AB | 3083 hCoV-19/England/MILK-CA9780/2020\|EPI_ISL_762251\|20 | ···················· | ···················· | ···················· | ···················· | ···················· | ···················· |
| AB | 3084 hCoV-19/England/MILK-CA95A4/2020\|EPI_ISL_762265\|20 | ···················· | ···················· | ···················· | ···················· | ···················· | ···················· |
| AB | 3085 hCoV-19/England/MILK-CA44F7/2020\|EPI_ISL_762267\|20 | ···················· | ···················· | ···················· | ···················· | ···················· | ···················· |
| AB | 3086 hCoV-19/England/MILK-CA9D51/2020\|EPI_ISL_762272\|20 | ···················· | ···················· | ···················· | ···················· | ···················· | ···················· |
| AB | 3087 hCoV-19/England/MILK-CA4A40/2020\|EPI_ISL_762273\|20 | ···················· | ·A·················· | ···················· | ···················· | ···················· | ···················· |
| AB | 3088 hCoV-19/England/MILK-CA4CC2/2020\|EPI_ISL_762275\|20 | ···················· | ···················· | ···················· | ···················· | ···················· | ···················· |
| AB | 3089 hCoV-19/England/MILK-CA372C/2020\|EPI_ISL_762281\|20 | ···················· | ···················· | ···················· | ···················· | ···················· | ···················· |
| AB | 3090 hCoV-19/England/MILK-CA3EAF/2020\|EPI_ISL_762284\|20 | ···················· | ···················· | ···················· | ···················· | ···················· | ···················· |
| AB | 3092 hCoV-19/Wales/PHWC-4A50CD/2020\|EPI_ISL_764819\|2020 | ···················· | ···················· | ···················· | ···················· | ···················· | ···················· |
| AB | 3094 hCoV-19/Wales/PHWC-4A5449/2020\|EPI_ISL_764866\|2020 | ···················· | ···················· | ···················· | ···················· | ···················· | ···················· |

| | | | | | | | | |
|---|---|---|---|---|---|---|---|---|
| AB | 3095 hCoV-19/Wales/PHWC-4A5458/2020\|EPI_ISL_764867\|2020 | ···················· | ···················· | ···················· | ···················· | ···················· | ···················· | |
| AB | 3096 hCoV-19/Wales/PHWC-4A5467/2020\|EPI_ISL_764868\|2020 | ···················· | ···················· | ···················· | ···················· | ···················· | ···················· | |
| AB | 3097 hCoV-19/Wales/PHWC-4A5476/2020\|EPI_ISL_764869\|2020 | ···················· | ···················· | ···················· | ···················· | ···················· | ···················· | |
| AB | 3098 hCoV-19/Wales/PHWC-4A5485/2020\|EPI_ISL_764870\|2020 | ···················· | ···················· | ···················· | ···················· | ···················· | ···················· | |
| AB | 3099 hCoV-19/Wales/PHWC-4A5494/2020\|EPI_ISL_764871\|2020 | ···················· | ···················· | ···················· | ···················· | ···················· | ···················· | |
| AB | 3100 hCoV-19/Wales/PHWC-4A54A3/2020\|EPI_ISL_764872\|2020 | ···················· | ···················· | ···················· | ···················· | ···················· | ···················· | |
| AB | 3101 hCoV-19/Wales/PHWC-4A54B2/2020\|EPI_ISL_764873\|2020 | ···················· | ···················· | ···················· | ···················· | ···················· | ···················· | |
| AB | 3102 hCoV-19/Wales/PHWC-4A54C1/2020\|EPI_ISL_764874\|2020 | ···················· | ···················· | ···················· | ···················· | ···················· | ···················· | |
| AB | 3103 hCoV-19/Wales/PHWC-4A54D0/2020\|EPI_ISL_764875\|2020 | ···················· | ···················· | ···················· | ···················· | ···················· | ···················· | |
| AB | 3104 hCoV-19/Wales/PHWC-4A54EF/2020\|EPI_ISL_764876\|2020 | ···················· | ···················· | ···················· | ···················· | ···················· | ···················· | |
| AB | 3105 hCoV-19/Wales/PHWC-4A54FE/2020\|EPI_ISL_764877\|2020 | ···················· | ···················· | ···················· | ···················· | ···················· | ···················· | |
| AB | 3107 hCoV-19/Wales/PHWC-4A5519/2020\|EPI_ISL_764879\|2020 | ···················· | ···················· | ···················· | ···················· | ···················· | ···················· | |
| AB | 3108 hCoV-19/Wales/PHWC-4A5528/2020\|EPI_ISL_764880\|2020 | ···················· | ···················· | ···················· | ···················· | ···················· | ···················· | |
| AB | 3109 hCoV-19/Wales/PHWC-4A5537/2020\|EPI_ISL_764881\|2020 | ···················· | ···················· | ···················· | ···················· | ···················· | ···················· | |
| AB | 3110 hCoV-19/Wales/PHWC-4A5546/2020\|EPI_ISL_764882\|2020 | ···················· | ···················· | ···················· | ···················· | ···················· | ···················· | |
| AB | 3111 hCoV-19/Wales/PHWC-4A5555/2020\|EPI_ISL_764883\|2020 | ···················· | ···················· | ···················· | ···················· | ···················· | ···················· | |
| AB | 3112 hCoV-19/Wales/PHWC-4A5564/2020\|EPI_ISL_764884\|2020 | ···················· | ···················· | ···················· | ···················· | ···················· | ···················· | |
| AB | 3113 hCoV-19/Wales/PHWC-4A5573/2020\|EPI_ISL_764885\|2020 | ···················· | ···················· | ···················· | ···················· | ···················· | ···················· | |
| AB | 3115 hCoV-19/Wales/PHWC-4A5591/2020\|EPI_ISL_764887\|2020 | ···················· | ···················· | ···················· | ···················· | ···················· | ···················· | |
| AB | 3116 hCoV-19/Wales/PHWC-4A55A0/2020\|EPI_ISL_764888\|2020 | ···················· | ···················· | ···················· | ···················· | ···················· | ···················· | |
| AB | 3117 hCoV-19/Wales/PHWC-4A55BF/2020\|EPI_ISL_764889\|2020 | ···················· | ···················· | ···················· | ···················· | ···················· | ···················· | |
| AB | 3119 hCoV-19/Wales/PHWC-4A55DD/2020\|EPI_ISL_764891\|2020 | ···················· | ···················· | ···················· | ···················· | ···················· | ···················· | |
| AB | 3120 hCoV-19/Wales/PHWC-4A55EC/2020\|EPI_ISL_764892\|2020 | ···················· | ···················· | ···················· | ···················· | ···················· | ···················· | |
| AB | 3121 hCoV-19/Wales/PHWC-4A55FB/2020\|EPI_ISL_764893\|2020 | ···················· | ···················· | ···················· | ···················· | ···················· | ···················· | |
| AB | 3122 hCoV-19/Wales/PHWC-4A5607/2020\|EPI_ISL_764894\|2020 | ···················· | ···················· | ···················· | ···················· | ···················· | ···················· | |
| AB | 3123 hCoV-19/Wales/PHWC-4A5616/2020\|EPI_ISL_764895\|2020 | ···················· | ···················· | ···················· | ···················· | ···················· | ···················· | |
| AB | 3124 hCoV-19/Wales/PHWC-4A5625/2020\|EPI_ISL_764896\|2020 | ···················· | ···················· | ···················· | ···················· | NNN················· | ···················· | 1 |
| AB | 3125 hCoV-19/Wales/PHWC-4A5643/2020\|EPI_ISL_764897\|2020 | ···················· | ···················· | ···················· | ···················· | ···················· | ···················· | |
| AB | 3126 hCoV-19/Wales/PHWC-4A5652/2020\|EPI_ISL_764898\|2020 | ···················· | ···················· | ···················· | ···················· | ···················· | ···················· | |
| AB | 3127 hCoV-19/Wales/PHWC-4A5661/2020\|EPI_ISL_764899\|2020 | ···················· | ···················· | ···················· | ···················· | ···················· | ···················· | |
| AB | 3128 hCoV-19/USA/LA-EVTL1131/2020\|EPI_ISL_768400\|2020-1 | ···················· | ···················· | ···················· | ···················· | ···················· | ···················· | |
| AB | 3129 hCoV-19/USA/LA-EVTL1132/2020\|EPI_ISL_768401\|2020-1 | ···················· | ·A·················· | ···················· | ···················· | ···················· | ···················· | |
| AB | 3130 hCoV-19/USA/LA-EVTL1133/2020\|EPI_ISL_768402\|2020-1 | ···················· | ·A·················· | ···················· | ···················· | ···················· | ···················· | |
| AB | 3131 hCoV-19/USA/LA-EVTL1134/2020\|EPI_ISL_768403\|2020-1 | ···················· | ·A·················· | ···················· | ···················· | ···················· | ···················· | |
| AB | 3132 hCoV-19/USA/LA-EVTL1135/2020\|EPI_ISL_768404\|2020-1 | ···················· | ···················· | ···················· | ···················· | ···················· | ···················· | |
| AB | 3133 hCoV-19/USA/LA-EVTL1137/2020\|EPI_ISL_768406\|2020-1 | ···················· | ···················· | ···················· | ···················· | ···················· | ···················· | |
| AB | 3134 hCoV-19/USA/LA-EVTL1138/2020\|EPI_ISL_768407\|2020-1 | ···················· | ···················· | ···················· | ···················· | ···················· | ···················· | |
| AB | 3135 hCoV-19/USA/LA-EVTL1139/2020\|EPI_ISL_768408\|2020-1 | ···················· | ···················· | ···················· | ···················· | ···················· | ···················· | |
| AB | 3136 hCoV-19/USA/LA-EVTL1140/2020\|EPI_ISL_768409\|2020-1 | ···················· | ···················· | ···················· | ···················· | ···················· | ···················· | |
| AB | 3137 hCoV-19/USA/LA-EVTL1141/2020\|EPI_ISL_768410\|2020-1 | ···················· | ···················· | ···················· | ···················· | ···················· | ···················· | |
| AB | 3138 hCoV-19/USA/LA-EVTL1142/2020\|EPI_ISL_768411\|2020-1 | ···················· | ·A·················· | ···················· | ···················· | ···················· | ···················· | |
| AB | 3139 hCoV-19/USA/LA-EVTL1143/2020\|EPI_ISL_768412\|2020-1 | ···················· | ·A·················· | ···················· | ···················· | ···················· | ···················· | |
| AB | 3140 hCoV-19/USA/LA-EVTL1144/2020\|EPI_ISL_768413\|2020-1 | ···················· | ···················· | ···················· | ···················· | ···················· | ···················· | |
| AB | 3141 hCoV-19/USA/LA-EVTL1145/2020\|EPI_ISL_768414\|2020-1 | ···················· | ···················· | ···················· | ···················· | ···················· | ···················· | |
| AB | 3142 hCoV-19/USA/LA-EVTL1146/2020\|EPI_ISL_768415\|2020-1 | ···················· | ·A·················· | ···················· | ···················· | ···················· | ···················· | |
| AB | 3143 hCoV-19/USA/LA-EVTL1147/2020\|EPI_ISL_768416\|2020-1 | ···················· | ···················· | ···················· | ···················· | ···················· | ···················· | |
| AB | 3144 hCoV-19/USA/LA-EVTL1148/2020\|EPI_ISL_768417\|2020-1 | ···················· | ·A·················· | ···················· | ···················· | ···················· | ···················· | |
| AB | 3145 hCoV-19/USA/LA-EVTL1152/2020\|EPI_ISL_768421\|2020-1 | ···················· | ···················· | ···················· | ···················· | ···················· | ···················· | |
| AB | 3146 hCoV-19/USA/LA-EVTL1153/2020\|EPI_ISL_768422\|2020-1 | ···················· | ···················· | ···················· | ···················· | ···················· | ···················· | |
| AB | 3147 hCoV-19/USA/LA-EVTL1154/2020\|EPI_ISL_768423\|2020-1 | ···················· | ···················· | ···················· | ···················· | ···················· | ···················· | |
| AB | 3148 hCoV-19/USA/LA-EVTL1155/2020\|EPI_ISL_768424\|2020-1 | ···················· | ···················· | ···················· | ···················· | ···················· | ···················· | |
| AB | 3149 hCoV-19/USA/LA-EVTL1156/2020\|EPI_ISL_768425\|2020-1 | ···················· | ·A·················· | ···················· | ···················· | ···················· | ···················· | |
| AB | 3150 hCoV-19/USA/LA-EVTL1157/2020\|EPI_ISL_768426\|2020-1 | ···················· | ·A·················· | ···················· | ···················· | ···················· | ···················· | |
| AB | 3151 hCoV-19/England/QEUH-C4BA78/2020\|EPI_ISL_769716\|20 | ···················· | ···················· | ···················· | ···················· | ···················· | ···················· | |
| AB | 3152 hCoV-19/England/QEUH-C4B7F9/2020\|EPI_ISL_769720\|20 | ···················· | ···················· | ···················· | ···················· | ···················· | ···················· | |
| AB | 3153 hCoV-19/England/MILK-C501D5/2020\|EPI_ISL_769723\|20 | ···················· | ···················· | ···················· | ···················· | ···················· | ···················· | |
| AB | 3154 hCoV-19/England/QEUH-C4B577/2020\|EPI_ISL_769724\|20 | ···················· | ···················· | ···················· | ···················· | ···················· | ···················· | |
| AB | 3155 hCoV-19/England/MILK-C53460/2020\|EPI_ISL_769728\|20 | ···················· | ···················· | ···················· | ···················· | ···················· | ···················· | |
| AB | 3156 hCoV-19/England/MILK-C53B10/2020\|EPI_ISL_769729\|20 | ···················· | ···················· | ···················· | ···················· | ···················· | ···················· | |
| AB | 3157 hCoV-19/England/MILK-C51611/2020\|EPI_ISL_769732\|20 | ···················· | ···················· | ···················· | ···················· | ···················· | ···················· | |
| AB | 3158 hCoV-19/England/MILK-C5171E/2020\|EPI_ISL_769733\|20 | ···················· | ···················· | ···················· | ···················· | ···················· | ···················· | |
| AB | 3159 hCoV-19/England/MILK-C5182A/2020\|EPI_ISL_769738\|20 | ···················· | ···················· | ···················· | ···················· | ···················· | ···················· | |
| AB | 3160 hCoV-19/England/QEUH-C4B61A/2020\|EPI_ISL_769743\|20 | ···················· | ···················· | ···················· | ···················· | ···················· | ···················· | |
| AB | 3161 hCoV-19/England/MILK-C517C3/2020\|EPI_ISL_769744\|20 | ···················· | ···················· | ···················· | ···················· | ···················· | ···················· | |
| AB | 3162 hCoV-19/England/MILK-C514F9/2020\|EPI_ISL_769746\|20 | ···················· | ···················· | ···················· | ···················· | ······T············· | ···················· | |
| AB | 3163 hCoV-19/England/MILK-C53855/2020\|EPI_ISL_769747\|20 | ···················· | ···················· | ···················· | ···················· | ···················· | ···················· | |
| AB | 3164 hCoV-19/England/MILK-C4FDC6/2020\|EPI_ISL_769748\|20 | ···················· | ···················· | ···················· | ···················· | ···················· | ···················· | |
| AB | 3165 hCoV-19/England/QEUH-C4B5E0/2020\|EPI_ISL_769752\|20 | ···················· | ···················· | ···················· | ···················· | ···················· | ···················· | |
| AB | 3166 hCoV-19/England/QEUH-C4B89C/2020\|EPI_ISL_769753\|20 | ···················· | ···················· | ···················· | ···················· | ···················· | ···················· | |
| AB | 3167 hCoV-19/England/QEUH-C4B683/2020\|EPI_ISL_769758\|20 | ···················· | ···················· | ···················· | ···················· | ···················· | ···················· | |
| AB | 3168 hCoV-19/Germany/NW-HHU-466/2020\|EPI_ISL_770700\|202 | ···················· | ···················· | ···················· | ···················· | ···················· | ···················· | |
| AB | 3169 hCoV-19/Germany/NW-HHU-467/2020\|EPI_ISL_770701\|202 | ···················· | ···················· | ···················· | ···················· | ···················· | ···················· | |
| AB | 3170 hCoV-19/Germany/NW-HHU-469/2020\|EPI_ISL_770703\|202 | ···················· | ···················· | ···················· | ···················· | ···················· | ···················· | |
| AB | 3171 hCoV-19/Germany/NW-HHU-470/2020\|EPI_ISL_770704\|202 | ···················· | ···················· | ···················· | ···················· | ···················· | ···················· | |
| AB | 3172 hCoV-19/Germany/NW-HHU-471/2020\|EPI_ISL_770705\|202 | ···················· | ···················· | ···················· | ···················· | ···················· | ···················· | |
| AB | 3173 hCoV-19/Germany/NW-HHU-472/2020\|EPI_ISL_770706\|202 | ···················· | ···················· | ···················· | ···················· | ···················· | ···················· | |
| AB | 3174 hCoV-19/Germany/NW-HHU-473/2020\|EPI_ISL_770707\|202 | ···················· | ···················· | ···················· | ···················· | ···················· | ···················· | |
| AB | 3175 hCoV-19/Germany/NW-HHU-474/2020\|EPI_ISL_770708\|202 | ···················· | ···················· | ···················· | ···················· | ···················· | ···················· | |
| AB | 3176 hCoV-19/Germany/NW-HHU-475/2020\|EPI_ISL_770709\|202 | ···················· | ···················· | ···················· | ···················· | ···················· | ···················· | |
| AB | 3177 hCoV-19/Germany/NW-HHU-476/2020\|EPI_ISL_770710\|202 | ······C············· | ···················· | ···················· | ···················· | ···················· | ···················· | |

| | | | | | | | | |
|---|---|---|---|---|---|---|---|---|
| AB | 3178 | hCoV-19/Germany/NW-HHU-477/2020\|EPI_ISL_770711\|202 | ···················· | ···················· | ···················· | ···················· | ···················· | ···················· |
| AB | 3179 | hCoV-19/Germany/NW-HHU-479/2020\|EPI_ISL_770713\|202 | ···················· | ···················· | ···················· | ···················· | ···················· | ···················· |
| AB | 3180 | hCoV-19/Germany/NW-HHU-481/2020\|EPI_ISL_770715\|202 | ···················· | ···················· | ···················· | ···················· | ···················· | ···················· |
| AB | 3181 | hCoV-19/Germany/NW-HHU-482/2020\|EPI_ISL_770716\|202 | ···················· | ···················· | ···················· | ···················· | ···················· | ···················· |
| AB | 3182 | hCoV-19/Germany/NW-HHU-483/2020\|EPI_ISL_770717\|202 | ···················· | ···················· | ···················· | ···················· | ···················· | ···················· |
| AB | 3183 | hCoV-19/Germany/NW-HHU-484/2020\|EPI_ISL_770718\|202 | ···················· | ···················· | ···················· | ···················· | ···················· | ···················· |
| AB | 3184 | hCoV-19/Germany/NW-HHU-485/2020\|EPI_ISL_770719\|202 | ···················· | ···················· | ···················· | ···················· | ···················· | ···················· |
| AB | 3185 | hCoV-19/Germany/NW-HHU-504/2020\|EPI_ISL_770720\|202 | ···················· | ···················· | ···················· | ···················· | ···················· | ···················· |
| AB | 3186 | hCoV-19/Germany/NW-HHU-505/2020\|EPI_ISL_770721\|202 | ···················· | ······A············· | ···················· | ···················· | ···················· | ···················· |
| AB | 3187 | hCoV-19/Germany/NW-HHU-507/2020\|EPI_ISL_770723\|202 | ···················· | ···················· | ···················· | ···················· | ···················· | ···················· |
| AB | 3188 | hCoV-19/Germany/NW-HHU-508/2020\|EPI_ISL_770724\|202 | ···················· | ······A············· | ···················· | ···················· | ···················· | ···················· |
| AB | 3189 | hCoV-19/Germany/NW-HHU-514/2020\|EPI_ISL_770730\|202 | ···················· | ···················· | ···T················ | ···················· | ···················· | ···················· |
| AB | 3190 | hCoV-19/Germany/NW-HHU-486/2020\|EPI_ISL_770735\|202 | ···················· | ···················· | ···················· | ···················· | ···················· | ···················· |
| AB | 3191 | hCoV-19/Germany/NW-HHU-489/2020\|EPI_ISL_770738\|202 | ···················· | ···················· | ···················· | ···················· | ···················· | ···················· |
| AB | 3192 | hCoV-19/Germany/NW-HHU-490/2020\|EPI_ISL_770739\|202 | ···················· | ···················· | ···················· | ···················· | ···················· | ···················· |
| AB | 3193 | hCoV-19/Norway/6693/2020\|EPI_ISL_775513\|2020-11-29 | ···················· | ···················· | ···················· | ···················· | ···················· | ··A················· |
| AB | 3194 | hCoV-19/Norway/6695/2020\|EPI_ISL_775514\|2020-11-18 | ···················· | ···················· | ···················· | ···················· | ···················· | ···················· |
| AB | 3195 | hCoV-19/Norway/6697/2020\|EPI_ISL_775515\|2020-11-13 | ···················· | ···················· | ···················· | ···················· | ···················· | ···················· |
| AB | 3196 | hCoV-19/Norway/6701/2020\|EPI_ISL_775516\|2020-12-07 | ···················· | ···················· | ···················· | ···················· | ···················· | ···················· |
| AB | 3197 | hCoV-19/Norway/6705/2020\|EPI_ISL_775517\|2020-12-07 | ···················· | ···················· | ···················· | ···················· | ···················· | ···················· |
| AB | 3198 | hCoV-19/Norway/6778/2020\|EPI_ISL_775518\|2020-12-12 | ···················· | ···················· | ···················· | ···················· | ···················· | ···················· |
| AB | 3199 | hCoV-19/Norway/6779/2020\|EPI_ISL_775519\|2020-12-10 | ···················· | ···················· | ···················· | ···················· | ···················· | ···················· |
| AB | 3200 | hCoV-19/Norway/6780/2020\|EPI_ISL_775520\|2020-12-09 | ···················· | ···················· | ···················· | ···················· | ···················· | ···················· |
| AB | 3201 | hCoV-19/Norway/6781/2020\|EPI_ISL_775521\|2020-12-10 | ···················· | ·A·················· | ···················· | ···················· | ···················· | ··A················· |
| AB | 3202 | hCoV-19/Norway/6782/2020\|EPI_ISL_775522\|2020-12-10 | ···················· | ···················· | ···················· | ···················· | ···················· | ···················· |
| AB | 3203 | hCoV-19/Norway/4036/2020\|EPI_ISL_775523\|2020-09-28 | ···················· | ···················· | ···················· | ···················· | ···················· | ···················· |
| AB | 3204 | hCoV-19/Norway/7104/2020\|EPI_ISL_775524\|2020-12-21 | ···················· | ·A·················· | ···················· | ···················· | ···················· | ··A················· |
| AB | 3205 | hCoV-19/Norway/7105/2020\|EPI_ISL_775525\|2020-12-21 | ···················· | ·A·················· | ···················· | ···················· | ···················· | ··A················· |
| AB | 3206 | hCoV-19/Norway/7106/2020\|EPI_ISL_775526\|2020-12-21 | ···················· | ·A·················· | ···················· | ···················· | ···················· | ··A················· |
| AB | 3207 | hCoV-19/Norway/7107/2020\|EPI_ISL_775527\|2020-12-21 | ···················· | ·A·················· | ···················· | ···················· | ···················· | ··A················· |
| AB | 3208 | hCoV-19/Norway/7023/2020\|EPI_ISL_775528\|2020-12-17 | ···················· | ···················· | ···················· | ···················· | ···················· | ···················· |
| AB | 3209 | hCoV-19/Norway/7024/2020\|EPI_ISL_775529\|2020-12-14 | ···················· | ···················· | ···················· | ···················· | ···················· | ···················· |
| AB | 3210 | hCoV-19/Norway/7025/2020\|EPI_ISL_775530\|2020-12-17 | ···················· | ···················· | ···················· | ···················· | ···················· | ···················· |
| AB | 3211 | hCoV-19/Sweden/20-09926/2020\|EPI_ISL_775531\|2020-1 | ···················· | ···················· | ···················· | ···················· | ···················· | ···················· |
| AB | 3212 | hCoV-19/Sweden/20-09927/2020\|EPI_ISL_775532\|2020-1 | ···················· | ···················· | ···················· | ···················· | ···················· | ···················· |
| AB | 3213 | hCoV-19/Sweden/20-09932/2020\|EPI_ISL_775533\|2020-1 | ···················· | ···················· | ···················· | ···················· | ···················· | ···················· |
| AB | 3214 | hCoV-19/Sweden/20-09935/2020\|EPI_ISL_775534\|2020-1 | ···················· | ···················· | ···················· | ···················· | ···················· | ···················· |
| AB | 3215 | hCoV-19/Sweden/20-09939/2020\|EPI_ISL_775535\|2020-1 | ···················· | ···················· | ···················· | ···················· | ···················· | ···················· |
| AB | 3216 | hCoV-19/Sweden/20-09940/2020\|EPI_ISL_775536\|2020-1 | ···················· | ···················· | ···················· | ···················· | ···················· | ···················· |
| AB | 3217 | hCoV-19/Sweden/20-09968/2020\|EPI_ISL_775537\|2020-1 | ···················· | ···················· | ···················· | ···················· | ···················· | ···················· |
| AB | 3218 | hCoV-19/Sweden/20-09970/2020\|EPI_ISL_775538\|2020-1 | ···················· | ···················· | ···················· | ···················· | ···················· | ···················· |
| AB | 3219 | hCoV-19/Sweden/20-09971/2020\|EPI_ISL_775539\|2020-1 | ···················· | ···················· | ···················· | ···················· | ···················· | ···················· |
| AB | 3220 | hCoV-19/Sweden/20-09974/2020\|EPI_ISL_775540\|2020-1 | ···················· | ···················· | ···················· | ···················· | ···················· | ···················· |
| AB | 3221 | hCoV-19/Sweden/20-09990/2020\|EPI_ISL_775541\|2020-1 | ···················· | ···················· | ···················· | ···················· | ···················· | ···················· |
| AB | 3222 | hCoV-19/Sweden/20-53713/2020\|EPI_ISL_775542\|2020-1 | ···················· | ···················· | ···················· | ···················· | ···················· | ···················· |
| AB | 3223 | hCoV-19/Sweden/20-53714/2020\|EPI_ISL_775543\|2020-1 | ···················· | ···················· | ···················· | ···················· | ···················· | ···················· |
| AB | 3224 | hCoV-19/Sweden/20-53721/2020\|EPI_ISL_775544\|2020-1 | ···················· | ···················· | ···················· | ···················· | ···················· | ···················· |
| AB | 3225 | hCoV-19/Sweden/20-53724/2020\|EPI_ISL_775545\|2020-1 | ···················· | ···················· | ···················· | ···················· | ······T············· | ···················· |
| AB | 3226 | hCoV-19/Sweden/20-53777/2020\|EPI_ISL_775546\|2020-1 | ···················· | ···················· | ···················· | ···················· | ···················· | ···················· |
| AB | 3227 | hCoV-19/Sweden/20-53783/2020\|EPI_ISL_775547\|2020-1 | ···················· | ···················· | ···················· | ···················· | ···················· | ···················· |
| AB | 3228 | hCoV-19/Sweden/20-53789/2020\|EPI_ISL_775548\|2020-1 | ···················· | ···················· | ···················· | ···················· | ···················· | ···················· |
| AB | 3229 | hCoV-19/Sweden/20-13018/2020\|EPI_ISL_775549\|2020-1 | ···················· | ···················· | ···················· | ···················· | ···················· | ···················· |
| AB | 3230 | hCoV-19/Sweden/20-13027/2020\|EPI_ISL_775550\|2020-1 | ···················· | ···················· | ···················· | ···················· | ···················· | ···················· |
| AB | 3231 | hCoV-19/Sweden/20-13029/2020\|EPI_ISL_775551\|2020-1 | ···················· | ···················· | ···················· | ···················· | ···················· | ···················· |
| AB | 3232 | hCoV-19/Sweden/20-13038/2020\|EPI_ISL_775552\|2020-1 | ···················· | ···················· | ···················· | ···················· | ···················· | ···················· |
| AB | 3233 | hCoV-19/Sweden/20-13040/2020\|EPI_ISL_775553\|2020-1 | ···················· | ···················· | ···················· | ···················· | ···················· | ···················· |
| AB | 3234 | hCoV-19/Sweden/20-13042/2020\|EPI_ISL_775554\|2020-1 | ···················· | ···················· | ···················· | ···················· | ···················· | ···················· |
| AB | 3235 | hCoV-19/Sweden/20-13046/2020\|EPI_ISL_775555\|2020-1 | ···················· | ···················· | ···················· | ···················· | ···················· | ···················· |
| AB | 3236 | hCoV-19/Sweden/20-13049/2020\|EPI_ISL_775556\|2020-1 | ···················· | ···················· | ···················· | ···················· | ···················· | ···················· |
| AB | 3237 | hCoV-19/Sweden/20-13061/2020\|EPI_ISL_775558\|2020-1 | ···················· | ···················· | ···················· | ···················· | ···················· | ···················· |
| AB | 3238 | hCoV-19/Sweden/20-13064/2020\|EPI_ISL_775559\|2020-1 | ···················· | ···················· | ···················· | ···················· | ···················· | ···················· |
| AB | 3239 | hCoV-19/Sweden/20-13071/2020\|EPI_ISL_775560\|2020-1 | ···················· | ···················· | ···················· | ···················· | ···················· | ···················· |
| AB | 3240 | hCoV-19/Sweden/20-13085/2020\|EPI_ISL_775561\|2020-1 | ···················· | ···················· | ···················· | ···················· | ···················· | ···················· |
| AB | 3241 | hCoV-19/Sweden/20-13094/2020\|EPI_ISL_775562\|2020-1 | ···················· | ···················· | ···················· | ···················· | ···················· | ···················· |
| AB | 3242 | hCoV-19/Sweden/20-13105/2020\|EPI_ISL_775564\|2020-1 | ···················· | ···················· | ···················· | ···················· | ···················· | ···················· |
| AB | 3243 | hCoV-19/Sweden/20-13106/2020\|EPI_ISL_775565\|2020-1 | ···················· | ···················· | ···················· | ···················· | ···················· | ···················· |
| AB | 3244 | hCoV-19/Sweden/20-13107/2020\|EPI_ISL_775566\|2020-1 | ···················· | ···················· | ···················· | ···················· | ···················· | ···················· |
| AB | 3245 | hCoV-19/Sweden/20-13110/2020\|EPI_ISL_775567\|2020-1 | ···················· | ···················· | ···················· | ···················· | ···················· | ···················· |
| AB | 3246 | hCoV-19/Sweden/20-13117/2020\|EPI_ISL_775568\|2020-1 | ···················· | ···················· | ···················· | ···················· | ···················· | ···················· |
| AB | 3247 | hCoV-19/Sweden/20-13120/2020\|EPI_ISL_775569\|2020-1 | ···················· | ···················· | ···················· | ···················· | ···················· | ···················· |
| AB | 3248 | hCoV-19/Sweden/20-13123/2020\|EPI_ISL_775570\|2020-1 | ···················· | ···················· | ···················· | ···················· | ···················· | ···················· |
| AB | 3249 | hCoV-19/Sweden/20-13124/2020\|EPI_ISL_775571\|2020-1 | ···················· | ···················· | ···················· | ···················· | ···················· | ···················· |
| AB | 3250 | hCoV-19/Sweden/20-13126/2020\|EPI_ISL_775572\|2020-1 | ···················· | ···················· | ···················· | ···················· | ···················· | ···················· |
| AB | 3251 | hCoV-19/Sweden/20-13129/2020\|EPI_ISL_775573\|2020-1 | ···················· | ···················· | ···················· | ···················· | ···················· | ···················· |
| AB | 3252 | hCoV-19/Sweden/20-13132/2020\|EPI_ISL_775574\|2020-1 | ···················· | ···················· | ···················· | ···················· | ···················· | ···················· |
| AB | 3253 | hCoV-19/Sweden/20-13134/2020\|EPI_ISL_775575\|2020-1 | ···················· | ···················· | ···················· | ···················· | ···················· | ···············A···· |
| AB | 3254 | hCoV-19/Sweden/20-13135/2020\|EPI_ISL_775576\|2020-1 | ···················· | ···················· | ···················· | ···················· | ···················· | ···················· |
| AB | 3255 | hCoV-19/Sweden/20-13136/2020\|EPI_ISL_775577\|2020-1 | ···················· | ···················· | ···················· | ···················· | ···················· | ···············A···· |
| AB | 3256 | hCoV-19/Sweden/20-13138/2020\|EPI_ISL_775578\|2020-1 | ···················· | ···················· | ···················· | ···················· | ···················· | ···················· |
| AB | 3257 | hCoV-19/Sweden/20-13141/2020\|EPI_ISL_775579\|2020-1 | ···················· | ···················· | ···················· | ···················· | ···················· | ···················· |

| | | | | | | | | |
|---|---|---|---|---|---|---|---|---|
| AB | 3258 | hCoV-19/Sweden/20-13146/2020\|EPI_ISL_775580\|2020-1 | ···················· | ···················· | ····················· | ···················· | ······T············· | ···················· |
| AB | 3259 | hCoV-19/Sweden/20-13147/2020\|EPI_ISL_775581\|2020-1 | ···················· | ···················· | ····················· | ···················· | ···················· | ···············A···· |
| AB | 3260 | hCoV-19/Sweden/20-53779/2020\|EPI_ISL_775583\|2020-1 | ···················· | ···················· | ····················· | ···················· | ···················· | ···················· |
| AB | 3261 | hCoV-19/Sweden/20-53806/2020\|EPI_ISL_775585\|2020-1 | ···················· | ···················· | ····················· | ···················· | ······T············· | ···················· |
| AB | 3262 | hCoV-19/Sweden/20-53807/2020\|EPI_ISL_775586\|2020-1 | ···················· | ···················· | ····················· | ···················· | ···················· | ···················· |
| AB | 3263 | hCoV-19/Sweden/20-53825/2020\|EPI_ISL_775587\|2020-1 | ···················· | ···················· | ····················· | ···················· | ···················· | ···················· |
| AB | 3264 | hCoV-19/Sweden/20-53843/2020\|EPI_ISL_775588\|2020-1 | ···················· | ···················· | ····················· | ···················· | ······T············· | ···················· |
| AB | 3265 | hCoV-19/Sweden/20-53844/2020\|EPI_ISL_775589\|2020-1 | ···················· | ···················· | ····················· | ···················· | ···················· | ···················· |
| AB | 3266 | hCoV-19/Sweden/20-53849/2020\|EPI_ISL_775590\|2020-1 | ···················· | ···················· | ····················· | ···················· | ·············T······ | ···················· |
| AB | 3267 | hCoV-19/Sweden/20-53852/2020\|EPI_ISL_775591\|2020-1 | ···················· | ·A·················· | ····················· | ···················· | ···················· | ···················· |
| AB | 3268 | hCoV-19/Sweden/20-53856/2020\|EPI_ISL_775592\|2020-1 | ···················· | ···················· | ····················· | ···················· | ···················· | ···················· |
| AB | 3269 | hCoV-19/Sweden/20-53870/2020\|EPI_ISL_775593\|2020-1 | ···················· | ···················· | ····················· | ···················· | ···················· | ···············A···· |
| AB | 3270 | hCoV-19/Indonesia/LA-NIHRD-PME8719/2020\|EPI_ISL_77 | ···················· | ···················· | ····················· | ···················· | ···················· | ···A················ |
| AB | 3271 | hCoV-19/Indonesia/BA-NIHRD-PME5833/2020\|EPI_ISL_77 | ···················· | ···················· | ····················· | ···················· | ···················· | ···················· |
| AB | 3272 | hCoV-19/Indonesia/PA-NIHRD-PME8510/2020\|EPI_ISL_77 | ···················· | ···················· | ···A················· | ···················· | ···················· | ···················· |
| AB | 3273 | hCoV-19/Indonesia/NB-NIHRD-PME1393/2020\|EPI_ISL_77 | ···················· | ···················· | ····················· | ···················· | ···················· | ···················· |
| AB | 3274 | hCoV-19/Israel/CVL-144-ngs/2020\|EPI_ISL_776844\|202 | ···················· | ···················· | ····················· | ···················· | ···················· | ···················· |
| AB | 3275 | hCoV-19/Israel/CVL-158-ngs/2020\|EPI_ISL_776845\|202 | ···················· | ·A·················· | ····················· | ···················· | ···················· | ···················· |
| AB | 3276 | hCoV-19/Israel/CVL-160-ngs/2020\|EPI_ISL_776846\|202 | ···················· | ···················· | ····················· | ···················· | ···················· | ···················· |
| AB | 3277 | hCoV-19/Israel/CVL-164-ngs/2020\|EPI_ISL_776847\|202 | ···················· | ···················· | ····················· | ···················· | ···················· | ···················· |
| AB | 3278 | hCoV-19/Israel/CVL-166-ngs/2020\|EPI_ISL_776848\|202 | ···················· | ···················· | ····················· | ···················· | ···················· | ···················· |
| AB | 3279 | hCoV-19/Israel/CVL-187-ngs/2020\|EPI_ISL_776849\|202 | ···················· | ···················· | ····················· | ···················· | ···················· | ···················· |
| AB | 3280 | hCoV-19/Israel/CVL-192-ngs/2020\|EPI_ISL_776850\|202 | ···················· | ·A·················· | ····················· | ···················· | ···················· | ···················· |
| AB | 3281 | hCoV-19/Israel/CVL-194-ngs/2020\|EPI_ISL_776851\|202 | ···················· | ···················· | ····················· | ···················· | ···················· | ···················· |
| AB | 3282 | hCoV-19/Israel/CVL-203-ngs/2020\|EPI_ISL_776852\|202 | ···················· | ···················· | ····················· | ···················· | ···················· | ···················· |
| AB | 3283 | hCoV-19/Israel/CVL-244-ngs/2020\|EPI_ISL_776853\|202 | ···················· | ···················· | ····················· | ···················· | ···················· | ···················· |
| AB | 3284 | hCoV-19/Israel/CVL-245-ngs/2020\|EPI_ISL_776854\|202 | ···················· | ···················· | ····················· | ···················· | ···················· | ···················· |
| AB | 3285 | hCoV-19/Israel/CVL-248-ngs/2020\|EPI_ISL_776855\|202 | ···················· | ···················· | ····················· | ···················· | ···················· | ···················· |
| AB | 3286 | hCoV-19/Israel/CVL-249-ngs/2020\|EPI_ISL_776856\|202 | ···················· | ···················· | ····················· | ···················· | ···················· | ···················· |
| AB | 3287 | hCoV-19/Israel/CVL-251-ngs/2020\|EPI_ISL_776857\|202 | ···················· | ···················· | ····················· | ···················· | ···················· | ···················· |
| AB | 3288 | hCoV-19/Israel/CVL-253-ngs/2020\|EPI_ISL_776858\|202 | ···················· | ···················· | ····················· | ···················· | ···················· | ···················· |
| AB | 3289 | hCoV-19/Israel/CVL-256-ngs/2020\|EPI_ISL_776859\|202 | ···················· | ···················· | ····················· | ···················· | ···················· | ···················· |
| AB | 3290 | hCoV-19/Israel/CVL-264-ngs/2020\|EPI_ISL_776860\|202 | ···················· | ·A·················· | ····················· | ···················· | ···················· | ···················· |
| AB | 3291 | hCoV-19/Israel/CVL-266-ngs/2020\|EPI_ISL_776861\|202 | ···················· | ···················· | ····················· | ···················· | ···················· | ···················· |
| AB | 3292 | hCoV-19/Israel/CVL-277-ngs/2020\|EPI_ISL_776862\|202 | ···················· | ···················· | ····················· | ···················· | ···················· | ···················· |
| AB | 3293 | hCoV-19/Israel/CVL-281-ngs/2020\|EPI_ISL_776863\|202 | ···················· | ···················· | ····················· | ···················· | ···················· | ···················· |
| AB | 3294 | hCoV-19/Israel/CVL-283-ngs/2020\|EPI_ISL_776864\|202 | ···················· | ···················· | ····················· | ···················· | ···················· | ···················· |
| AB | 3295 | hCoV-19/Israel/CVL-284-ngs/2020\|EPI_ISL_776865\|202 | ···················· | ···················· | ····················· | ···················· | ···················· | ···················· |
| AB | 3296 | hCoV-19/Israel/CVL-286-ngs/2020\|EPI_ISL_776866\|202 | ···················· | ···················· | ····················· | ···················· | ···················· | ···················· |
| AB | 3297 | hCoV-19/Israel/CVL-288-ngs/2020\|EPI_ISL_776867\|202 | ···················· | ···················· | ····················· | ···················· | ···················· | ···················· |
| AB | 3298 | hCoV-19/Israel/CVL-455-ngs/2020\|EPI_ISL_776868\|202 | ···················· | ···················· | ····················· | ···················· | ···················· | ···················· |
| AB | 3299 | hCoV-19/USA/OH-ODH-SC35549/2020\|EPI_ISL_776869\|202 | ···················· | ···················· | ····················· | ···················· | ···················· | ···················· |
| AB | 3300 | hCoV-19/Spain/M-20230996/2020\|EPI_ISL_776870\|2020- | ···················· | ···················· | ····················· | ···················· | ················N··· | ···················· |
| AB | 3302 | hCoV-19/USA/OH-ODH-SC35535/2020\|EPI_ISL_776872\|202 | ···················· | ···················· | ····················· | ···················· | ···················· | ···················· |
| AB | 3303 | hCoV-19/USA/OH-ODH-SC35519/2020\|EPI_ISL_776874\|202 | ···················· | ···················· | ····················· | ···················· | ···················· | ···················· |
| AB | 3305 | hCoV-19/USA/OH-ODH-SC35469/2020\|EPI_ISL_776876\|202 | ···················· | ···················· | ····················· | ···················· | ···················· | ···················· |
| AB | 3306 | hCoV-19/Spain/M-20166261/2020\|EPI_ISL_776877\|2020- | ···················· | ···················· | ····················· | ···················· | ···················· | ···················· |
| AB | 3307 | hCoV-19/Italy/CAM-TIGEM-196/2020\|EPI_ISL_776879\|20 | ···················· | ···················· | ····················· | ···················· | ······T············· | ···················· |
| AB | 3308 | hCoV-19/Italy/CAM-TIGEM-202/2020\|EPI_ISL_776880\|20 | ···················· | ···················· | ····················· | ···················· | ···················· | ···················· |
| AB | 3309 | hCoV-19/Italy/CAM-TIGEM-200/2020\|EPI_ISL_776881\|20 | ···················· | ···················· | ····················· | ···················· | ···················· | ···················· |
| AB | 3310 | hCoV-19/Italy/CAM-TIGEM-195/2020\|EPI_ISL_776882\|20 | ···················· | ···················· | ····················· | ···················· | ···················· | ···················· |
| AB | 3311 | hCoV-19/Italy/CAM-TIGEM-199/2020\|EPI_ISL_776883\|20 | ···················· | ···················· | ····················· | ···················· | ···················· | ···················· |
| AB | 3312 | hCoV-19/Italy/CAM-TIGEM-194/2020\|EPI_ISL_776884\|20 | ···················· | ···················· | ····················· | ···················· | ···················· | ···················· |
| AB | 3313 | hCoV-19/Italy/CAM-TIGEM-219/2020\|EPI_ISL_776885\|20 | ···················· | ···················· | ····················· | ···················· | ···················· | ···················· |
| AB | 3314 | hCoV-19/Italy/CAM-TIGEM-207/2020\|EPI_ISL_776886\|20 | ···················· | ···················· | ····················· | ···················· | ···················· | ···················· |
| AB | 3315 | hCoV-19/Italy/CAM-TIGEM-230/2020\|EPI_ISL_776887\|20 | ···················· | ···················· | ····················· | ···················· | ···················· | ···················· |
| AB | 3316 | hCoV-19/Italy/CAM-TIGEM-208/2020\|EPI_ISL_776888\|20 | ···················· | ···················· | ····················· | ···················· | ···················· | ···················· |
| AB | 3317 | hCoV-19/Italy/CAM-TIGEM-211/2020\|EPI_ISL_776889\|20 | ···················· | ···················· | ····················· | ···················· | ···················· | ···················· |
| AB | 3318 | hCoV-19/Italy/CAM-TIGEM-210/2020\|EPI_ISL_776890\|20 | ···················· | ···················· | ····················· | ···················· | ···················· | ···················· |
| AB | 3319 | hCoV-19/Italy/CAM-TIGEM-216/2020\|EPI_ISL_776891\|20 | ···················· | ···················· | ····················· | ···················· | ···················· | ···················· |
| AB | 3320 | hCoV-19/Italy/CAM-TIGEM-225/2020\|EPI_ISL_776892\|20 | ···················· | ···················· | ····················· | ···················· | ···················· | ···················· |
| AB | 3321 | hCoV-19/Italy/CAM-TIGEM-233/2020\|EPI_ISL_776893\|20 | ···················· | ···················· | ····················· | ···················· | ···················· | ···················· |
| AB | 3322 | hCoV-19/Italy/CAM-TIGEM-231/2020\|EPI_ISL_776894\|20 | ···················· | ·A·················· | ····················· | ···················· | ···················· | ···················· |
| AB | 3323 | hCoV-19/Italy/CAM-TIGEM-248/2020\|EPI_ISL_776895\|20 | ···················· | ···················· | ····················· | ···················· | ···················· | ···················· |
| AB | 3324 | hCoV-19/Italy/CAM-TIGEM-247/2020\|EPI_ISL_776896\|20 | ···················· | ·A·················· | ····················· | ···················· | ···················· | ···················· |
| AB | 3325 | hCoV-19/Italy/CAM-TIGEM-229/2020\|EPI_ISL_776897\|20 | ···················· | ···················· | ····················· | ···················· | ···················· | ···················· |
| AB | 3326 | hCoV-19/Italy/CAM-TIGEM-239/2020\|EPI_ISL_776898\|20 | ···················· | ···················· | ····················· | ···················· | ···················· | ···················· |
| AB | 3327 | hCoV-19/Italy/CAM-TIGEM-236/2020\|EPI_ISL_776899\|20 | ···················· | ···················· | ····················· | ···················· | ···················· | ···················· |
| AB | 3328 | hCoV-19/USA/LA-EVTL1403/2020\|EPI_ISL_779069\|2020-1 | ···················· | ···················· | ····················· | ···················· | ···················· | ···················· |
| AB | 3330 | hCoV-19/Wales/PHWC-4A59A4/2020\|EPI_ISL_764943\|2020 | ···················· | ···················· | ····················· | ···················· | ···················· | ···················· |
| AB | 3333 | hCoV-19/Wales/PHWC-4A59D1/2020\|EPI_ISL_764946\|2020 | ···················· | ···················· | ····················· | ···················· | ···················· | ···················· |
| AB | 3334 | hCoV-19/Wales/PHWC-4A5A0B/2020\|EPI_ISL_764947\|2020 | ···················· | ···················· | ····················· | ···················· | ···················· | ···················· |
| AB | 3336 | hCoV-19/Wales/PHWC-4A5A38/2020\|EPI_ISL_764949\|2020 | ···················· | ···················· | ····················· | ···················· | ···················· | ···················· |
| AB | 3337 | hCoV-19/Wales/PHWC-4A5A56/2020\|EPI_ISL_764950\|2020 | ···················· | ···················· | ····················· | ···················· | ···················· | ···················· |
| AB | 3340 | hCoV-19/England/ALDP-CB5059/2020\|EPI_ISL_767102\|20 | ···················· | ···················· | ····················· | ···················· | ···················· | ···················· |
| AB | 3341 | hCoV-19/England/ALDP-CB6708/2020\|EPI_ISL_767103\|20 | ···················· | ···················· | ····················· | ···················· | ···················· | ···················· |
| AB | 3342 | hCoV-19/England/ALDP-CB5AB5/2020\|EPI_ISL_767104\|20 | ···················· | ···················· | ····················· | ···················· | ···················· | ···················· |
| AB | 3343 | hCoV-19/England/ALDP-CB55D2/2020\|EPI_ISL_767105\|20 | ···················· | ···················· | ····················· | ···················· | ···················· | ···················· |
| AB | 3344 | hCoV-19/England/ALDP-CB6A4B/2020\|EPI_ISL_767106\|20 | ···················· | ···················· | ····················· | ···················· | ···················· | ···················· |
| AB | 3345 | hCoV-19/England/MILK-CA92DA/2020\|EPI_ISL_767107\|20 | ···················· | ·A·················· | ····················· | ···················· | ···················· | ···A················ |

| | | | | | | | |
|---|---|---|---|---|---|---|---|
| AB | 3346 hCoV-19/England/ALDP-C88699/2020\|EPI_ISL_767116\|20 | ···················· | ···················· | ···················· | ···················· | ······T············· | ···················· |
| AB | 3347 hCoV-19/Wales/ALDP-C57FF2/2020\|EPI_ISL_767120\|2020 | ···················· | ···················· | ···················· | ···················· | ···················· | ···················· |
| AB | 3348 hCoV-19/Wales/ALDP-C57EC8/2020\|EPI_ISL_767121\|2020 | ···················· | ···················· | ·········A·········· | ···················· | ···················· | ···················· |
| AB | 3349 hCoV-19/Wales/ALDP-C5979E/2020\|EPI_ISL_767122\|2020 | ···················· | ···················· | ···················· | ···················· | ···················· | ···················· |
| AB | 3350 hCoV-19/Wales/ALDP-C57DAD/2020\|EPI_ISL_767123\|2020 | ···················· | ···················· | ···················· | ···················· | ···················· | ···················· |
| AB | 3351 hCoV-19/Wales/ALDP-C596FB/2020\|EPI_ISL_767124\|2020 | ···················· | ···················· | ···················· | ···················· | ···················· | ···················· |
| AB | 3352 hCoV-19/Wales/ALDP-C597BC/2020\|EPI_ISL_767127\|2020 | ···················· | ···················· | ···················· | ···················· | ···················· | ···················· |
| AB | 3353 hCoV-19/Wales/ALDP-C588F6/2020\|EPI_ISL_767129\|2020 | ···················· | ···················· | ···················· | ···················· | ···················· | ···················· |
| AB | 3354 hCoV-19/Wales/ALDP-C5A029/2020\|EPI_ISL_767130\|2020 | ···················· | ···················· | ···················· | ···················· | ···················· | ···················· |
| AB | 3355 hCoV-19/Wales/ALDP-C58049/2020\|EPI_ISL_767131\|2020 | ···················· | ···················· | ···················· | ···················· | ···················· | ···················· |
| AB | 3356 hCoV-19/Wales/ALDP-C58708/2020\|EPI_ISL_767132\|2020 | ···················· | ···················· | ···················· | ···················· | ···················· | ···················· |
| AB | 3357 hCoV-19/Wales/ALDP-C57EAA/2020\|EPI_ISL_767133\|2020 | ···················· | ···················· | ···················· | ···················· | ···················· | ···················· |
| AB | 3358 hCoV-19/Wales/MILK-C53DCF/2020\|EPI_ISL_767134\|2020 | ···················· | ···················· | ···················· | ···················· | ···················· | ···················· |
| AB | 3359 hCoV-19/Wales/ALDP-C48871/2020\|EPI_ISL_767135\|2020 | ···················· | ···················· | ···················· | ···················· | ···················· | ···················· |
| AB | 3360 hCoV-19/Wales/ALDP-C48880/2020\|EPI_ISL_767136\|2020 | ···················· | ···················· | ···················· | ···················· | ···················· | ···················· |
| AB | 3361 hCoV-19/Wales/CAMC-C403F0/2020\|EPI_ISL_767138\|2020 | ···················· | ···················· | ···················· | ···················· | ···················· | ···················· |
| AB | 3362 hCoV-19/Wales/ALDP-C47D64/2020\|EPI_ISL_767140\|2020 | ···················· | ···················· | ···················· | ···················· | ···················· | ···················· |
| AB | 3363 hCoV-19/Wales/ALDP-C49931/2020\|EPI_ISL_767141\|2020 | ···················· | ······A············· | ···················· | ···················· | ···················· | ···················· |
| AB | 3364 hCoV-19/Wales/ALDP-C47F6E/2020\|EPI_ISL_767142\|2020 | ···················· | ···················· | ···················· | ···················· | ···················· | ···················· |
| AB | 3365 hCoV-19/Wales/ALDP-C47FC8/2020\|EPI_ISL_767144\|2020 | ···················· | ···················· | ···················· | ···················· | ···················· | ···················· |
| AB | 3366 hCoV-19/Wales/ALDP-C497EC/2020\|EPI_ISL_767146\|2020 | ···················· | ···················· | ···················· | ···················· | ···················· | ···················· |
| AB | 3367 hCoV-19/Wales/ALDP-C4995F/2020\|EPI_ISL_767147\|2020 | ···················· | ······A············· | ···················· | ···················· | ···················· | ···················· |
| AB | 3368 hCoV-19/Wales/ALDP-C499D7/2020\|EPI_ISL_767148\|2020 | ···················· | ···················· | ···················· | ···················· | ···················· | ···················· |
| AB | 3369 hCoV-19/Wales/ALDP-C498DA/2020\|EPI_ISL_767150\|2020 | ···················· | ···················· | ···················· | ···················· | ···················· | ···················· |
| AB | 3370 hCoV-19/Wales/ALDP-C3B3F8/2020\|EPI_ISL_767151\|2020 | ···················· | ···················· | ···················· | ···················· | ···················· | ···················· |
| AB | 3371 hCoV-19/Wales/ALDP-C3A55D/2020\|EPI_ISL_767152\|2020 | ···················· | ···················· | ···················· | ···················· | ···················· | ···················· |
| AB | 3372 hCoV-19/Wales/ALDP-C3B3E9/2020\|EPI_ISL_767154\|2020 | ···················· | ···················· | ···················· | ···················· | ···················· | ···················· |
| AB | 3373 hCoV-19/Wales/ALDP-C3AE8F/2020\|EPI_ISL_767156\|2020 | ···················· | ···················· | ···················· | ···················· | ···················· | ···················· |
| AB | 3374 hCoV-19/Wales/ALDP-C39925/2020\|EPI_ISL_767157\|2020 | ···················· | ···················· | ···················· | ···················· | ···················· | ···················· |
| AB | 3375 hCoV-19/Wales/ALDP-C39961/2020\|EPI_ISL_767158\|2020 | ···················· | ···················· | ···················· | ···················· | ···················· | ···················· |
| AB | 3376 hCoV-19/Wales/ALDP-C3ADEC/2020\|EPI_ISL_767159\|2020 | ···················· | ···················· | ···················· | ···················· | ···················· | ···················· |
| AB | 3377 hCoV-19/Wales/ALDP-C3AE61/2020\|EPI_ISL_767160\|2020 | ···················· | ···················· | ···················· | ···················· | ···················· | ···················· |
| AB | 3378 hCoV-19/Wales/ALDP-C39916/2020\|EPI_ISL_767161\|2020 | ···················· | ···················· | ···················· | ···················· | ···················· | ···················· |
| AB | 3379 hCoV-19/Wales/ALDP-C39794/2020\|EPI_ISL_767162\|2020 | ···················· | ···················· | ···················· | ···················· | ···················· | ···················· |
| AB | 3380 hCoV-19/Wales/ALDP-C39BA7/2020\|EPI_ISL_767163\|2020 | ···················· | ···················· | ···················· | ···················· | ···················· | ···················· |
| AB | 3381 hCoV-19/Wales/ALDP-C3999E/2020\|EPI_ISL_767164\|2020 | ···················· | ···················· | ···················· | ···················· | ···················· | ···················· |
| AB | 3382 hCoV-19/Wales/ALDP-C3AE07/2020\|EPI_ISL_767166\|2020 | ···················· | ···················· | ···················· | ···················· | ···················· | ···················· |
| AB | 3383 hCoV-19/Wales/ALDP-C3B343/2020\|EPI_ISL_767167\|2020 | ···················· | ···················· | ···················· | ···················· | ···················· | ···················· |
| AB | 3384 hCoV-19/Wales/ALDP-C3B38F/2020\|EPI_ISL_767168\|2020 | ···················· | ···················· | ···················· | ···················· | ···················· | ···················· |
| AB | 3385 hCoV-19/Wales/ALDP-C3B3DA/2020\|EPI_ISL_767169\|2020 | ···················· | ···················· | ···················· | ···················· | ···················· | ···················· |
| AB | 3386 hCoV-19/Wales/ALDP-C3B413/2020\|EPI_ISL_767170\|2020 | ···················· | ···················· | ···················· | ···················· | ···················· | ···················· |
| AB | 3387 hCoV-19/Wales/ALDP-C39D29/2020\|EPI_ISL_767171\|2020 | ···················· | ······A············· | ···················· | ···················· | ···················· | ···················· |
| AB | 3388 hCoV-19/Wales/ALDP-C3AE9E/2020\|EPI_ISL_767172\|2020 | ···················· | ···················· | ···················· | ···················· | ···················· | ···················· |
| AB | 3389 hCoV-19/Wales/ALDP-C3A4E7/2020\|EPI_ISL_767173\|2020 | ···················· | ···················· | ···················· | ···················· | ···················· | ···················· |
| AB | 3390 hCoV-19/Wales/ALDP-C399DA/2020\|EPI_ISL_767174\|2020 | ···················· | ···················· | ···················· | ···················· | ···················· | ···················· |
| AB | 3391 hCoV-19/Wales/ALDP-C399F8/2020\|EPI_ISL_767175\|2020 | ···················· | ···················· | ···················· | ···················· | ···················· | ···················· |
| AB | 3392 hCoV-19/Wales/ALDP-C396D3/2020\|EPI_ISL_767176\|2020 | ···················· | ···················· | ···················· | ···················· | ······T············· | ···················· |
| AB | 3393 hCoV-19/Wales/ALDP-C39D47/2020\|EPI_ISL_767177\|2020 | ···················· | ···················· | ···················· | ···················· | ···················· | ···················· |
| AB | 3394 hCoV-19/Wales/ALDP-C3D62A/2020\|EPI_ISL_767178\|2020 | ···················· | ···················· | ···················· | ···················· | ···················· | ···················· |
| AB | 3395 hCoV-19/Wales/ALDP-C3B39E/2020\|EPI_ISL_767179\|2020 | ···················· | ···················· | ···················· | ···················· | ···················· | ···················· |
| AB | 3396 hCoV-19/USA/LA-EVTL1149/2020\|EPI_ISL_768418\|2020-1 | ···················· | ···················· | ···················· | ···················· | ···················· | ···················· |
| AB | 3397 hCoV-19/USA/LA-EVTL1150/2020\|EPI_ISL_768419\|2020-1 | ···················· | ···················· | ···················· | ···················· | ···················· | ···················· |
| AB | 3398 hCoV-19/USA/LA-EVTL1151/2020\|EPI_ISL_768420\|2020-1 | ···················· | ···················· | ···················· | ···················· | ···················· | ···················· |
| AB | 3399 hCoV-19/USA/LA-EVTL1158/2020\|EPI_ISL_768427\|2020-1 | ···················· | ·A·················· | ···················· | ···················· | ···················· | ···················· |
| AB | 3400 hCoV-19/USA/LA-EVTL1159/2020\|EPI_ISL_768428\|2020-1 | ···················· | ·A·················· | ···················· | ···················· | ···················· | ···················· |
| AB | 3401 hCoV-19/USA/LA-EVTL1160/2020\|EPI_ISL_768429\|2020-1 | ···················· | ·A·················· | ···················· | ···················· | ···················· | ···················· |
| AB | 3402 hCoV-19/USA/LA-EVTL1161/2020\|EPI_ISL_768430\|2020-1 | ···················· | ·A·················· | ···················· | ···················· | ···················· | ···················· |
| AB | 3403 hCoV-19/USA/LA-EVTL1162/2020\|EPI_ISL_768431\|2020-1 | ···················· | ·A·················· | ···················· | ···················· | ···················· | ···················· |
| AB | 3404 hCoV-19/USA/LA-EVTL1163/2020\|EPI_ISL_768432\|2020-1 | ···················· | ·A·················· | ···················· | ···················· | ···················· | ···················· |
| AB | 3405 hCoV-19/USA/LA-EVTL1164/2020\|EPI_ISL_768433\|2020-1 | ···················· | ·A·················· | ···················· | ···················· | ···················· | ···················· |
| AB | 3406 hCoV-19/USA/LA-EVTL1165/2020\|EPI_ISL_768434\|2020-1 | ···················· | ·A·················· | ···················· | ···················· | ···················· | ···················· |
| AB | 3407 hCoV-19/USA/LA-EVTL1166/2020\|EPI_ISL_768435\|2020-1 | ···················· | ·A·················· | ···················· | ···················· | ···················· | ···················· |
| AB | 3408 hCoV-19/USA/LA-EVTL1167/2020\|EPI_ISL_768436\|2020-1 | ···················· | ·A·················· | ···················· | ···················· | ···················· | ···················· |
| AB | 3409 hCoV-19/USA/LA-EVTL1168/2020\|EPI_ISL_768437\|2020-1 | ···················· | ·A·················· | ···················· | ···················· | ···················· | ···················· |
| AB | 3410 hCoV-19/USA/LA-EVTL1169/2020\|EPI_ISL_768438\|2020-1 | ···················· | ·A·················· | ···················· | ···················· | ···················· | ···················· |
| AB | 3411 hCoV-19/USA/LA-EVTL1170/2020\|EPI_ISL_768439\|2020-1 | ···················· | ···················· | ···················· | ···················· | ···················· | ···················· |
| AB | 3412 hCoV-19/USA/LA-EVTL1171/2020\|EPI_ISL_768440\|2020-1 | ···················· | ···················· | ···················· | ···················· | ···················· | ···················· |
| AB | 3413 hCoV-19/USA/LA-EVTL1172/2020\|EPI_ISL_768441\|2020-1 | ···················· | ···················· | ···················· | ···················· | ···················· | ···················· |
| AB | 3414 hCoV-19/USA/LA-EVTL1173/2020\|EPI_ISL_768442\|2020-1 | ···················· | ···················· | ···················· | ···················· | ···················· | ···················· |
| AB | 3415 hCoV-19/USA/LA-EVTL1174/2020\|EPI_ISL_768443\|2020-1 | ···················· | ···················· | ···················· | ···················· | ···················· | ···················· |
| AB | 3416 hCoV-19/USA/LA-EVTL1175/2020\|EPI_ISL_768444\|2020-1 | ···················· | ···················· | ···················· | ···················· | ···················· | ···················· |
| AB | 3417 hCoV-19/USA/LA-EVTL1176/2020\|EPI_ISL_768445\|2020-1 | ···················· | ···················· | ···················· | ···················· | ···················· | ···················· |
| AB | 3418 hCoV-19/USA/LA-EVTL1177/2020\|EPI_ISL_768446\|2020-1 | ···················· | ···················· | ···················· | ···················· | ···················· | ···················· |
| AB | 3419 hCoV-19/USA/LA-EVTL1178/2020\|EPI_ISL_768447\|2020-1 | ···················· | ···················· | ···················· | ···················· | ···················· | ···················· |
| AB | 3420 hCoV-19/USA/LA-EVTL1179/2020\|EPI_ISL_768448\|2020-1 | ···················· | ···················· | ···················· | ···················· | ···················· | ···················· |
| AB | 3421 hCoV-19/USA/LA-EVTL1180/2020\|EPI_ISL_768449\|2020-1 | ···················· | ···················· | ···················· | ···················· | ···················· | ···················· |
| AB | 3422 hCoV-19/USA/LA-EVTL1181/2020\|EPI_ISL_768450\|2020-1 | ···················· | ···················· | ···················· | ···················· | ···················· | ···················· |
| AB | 3423 hCoV-19/USA/LA-EVTL1182/2020\|EPI_ISL_768451\|2020-1 | ···················· | ···················· | ···················· | ···················· | ···················· | ···················· |
| AB | 3424 hCoV-19/USA/LA-EVTL1183/2020\|EPI_ISL_768452\|2020-1 | ···················· | ···················· | ···················· | ···················· | ···················· | ···················· |
| AB | 3425 hCoV-19/USA/LA-EVTL1184/2020\|EPI_ISL_768453\|2020-1 | ···················· | ···················· | ···················· | ···················· | ···················· | ···················· |

| | | | | | | | |
|---|---|---|---|---|---|---|---|
| AB | 3426 hCoV-19/USA/LA-EVTL1185/2020\|EPI_ISL_768454\|2020-1 | .................... | .................... | .................... | .................... | .................... | .................... |
| AB | 3427 hCoV-19/USA/LA-EVTL1186/2020\|EPI_ISL_768455\|2020-1 | .................... | .A.................. | .................... | .................... | .................... | .................... |
| AB | 3428 hCoV-19/USA/LA-EVTL1187/2020\|EPI_ISL_768456\|2020-1 | .................... | .................... | .................... | .................... | .................... | .................... |
| AB | 3429 hCoV-19/USA/LA-EVTL1188/2020\|EPI_ISL_768457\|2020-1 | .................... | .................... | .................... | .................... | .................... | .................... |
| AB | 3430 hCoV-19/USA/LA-EVTL1189/2020\|EPI_ISL_768458\|2020-1 | .................... | .................... | .................... | .................... | .................... | .................... |
| AB | 3431 hCoV-19/USA/LA-EVTL1190/2020\|EPI_ISL_768459\|2020-1 | .................... | .................... | .................... | .................... | .................... | .................... |
| AB | 3432 hCoV-19/USA/LA-EVTL1191/2020\|EPI_ISL_768460\|2020-1 | .................... | .................... | .................... | .................... | .................... | .................... |
| AB | 3433 hCoV-19/USA/LA-EVTL1192/2020\|EPI_ISL_768461\|2020-1 | .................... | .A.................. | .................... | .................... | .................... | .................... |
| AB | 3434 hCoV-19/USA/LA-EVTL1193/2020\|EPI_ISL_768462\|2020-1 | .................... | .................... | .................... | .................... | .................... | .................... |
| AB | 3435 hCoV-19/USA/LA-EVTL1194/2020\|EPI_ISL_768463\|2020-1 | .................... | .................... | .................... | .................... | .................... | .................... |
| AB | 3436 hCoV-19/USA/LA-EVTL1195/2020\|EPI_ISL_768464\|2020-1 | .................... | .................... | .................... | .................... | .................... | .................... |
| AB | 3437 hCoV-19/USA/LA-EVTL1196/2020\|EPI_ISL_768465\|2020-1 | .................... | .................... | .................... | .................... | .................... | .................... |
| AB | 3438 hCoV-19/USA/LA-EVTL1197/2020\|EPI_ISL_768466\|2020-1 | .................... | .................... | .................... | .................... | .................... | .................... |
| AB | 3439 hCoV-19/USA/LA-EVTL1198/2020\|EPI_ISL_768467\|2020-1 | .................... | .................... | .................... | .................... | .................... | .................... |
| AB | 3440 hCoV-19/USA/LA-EVTL1199/2020\|EPI_ISL_768468\|2020-1 | .................... | .................... | .................... | .................... | .................... | .................... |
| AB | 3441 hCoV-19/USA/LA-EVTL1200/2020\|EPI_ISL_768469\|2020-1 | .................... | .................... | .................... | .................... | .................... | .................... |
| AB | 3442 hCoV-19/USA/LA-EVTL1201/2020\|EPI_ISL_768470\|2020-1 | .................... | .A.................. | .................... | .................... | .................... | .................... |
| AB | 3443 hCoV-19/USA/LA-EVTL1202/2020\|EPI_ISL_768471\|2020-1 | .................... | .................... | .................... | .................... | .................... | .................... |
| AB | 3444 hCoV-19/USA/LA-EVTL1203/2020\|EPI_ISL_768472\|2020-1 | .................... | .................... | .................... | .................... | .................... | .................... |
| AB | 3445 hCoV-19/USA/LA-EVTL1206/2020\|EPI_ISL_768475\|2020-1 | .................... | .A.................. | .................... | .................... | .................... | .................... |
| AB | 3446 hCoV-19/USA/LA-EVTL1207/2020\|EPI_ISL_768476\|2020-1 | .................... | .................... | .................... | .................... | .................... | .................... |
| AB | 3447 hCoV-19/USA/LA-EVTL1208/2020\|EPI_ISL_768477\|2020-1 | .................... | .A.................. | .................... | .................... | .................... | .................... |
| AB | 3448 hCoV-19/USA/LA-EVTL1209/2020\|EPI_ISL_768478\|2020-1 | .................... | .................... | .................... | .................... | .................... | .................... |
| AB | 3449 hCoV-19/USA/LA-EVTL1210/2020\|EPI_ISL_768479\|2020-1 | .................... | .A.................. | .................... | .................... | .................... | .................... |
| AB | 3450 hCoV-19/USA/LA-EVTL1211/2020\|EPI_ISL_768480\|2020-1 | .................... | .................... | .................... | .................... | .................... | .................... |
| AB | 3451 hCoV-19/USA/LA-EVTL1212/2020\|EPI_ISL_768481\|2020-1 | .................... | .................... | .................... | .................... | .................... | .................... |
| AB | 3452 hCoV-19/USA/LA-EVTL1213/2020\|EPI_ISL_768482\|2020-1 | .................... | .A.................. | .................... | .................... | .................... | .................... |
| AB | 3453 hCoV-19/USA/LA-EVTL1214/2020\|EPI_ISL_768483\|2020-1 | .................... | .................... | .................... | .................... | .................... | .................... |
| AB | 3454 hCoV-19/USA/LA-EVTL1215/2020\|EPI_ISL_768484\|2020-1 | .................... | .................... | .................... | .................... | .................... | .................... |
| AB | 3455 hCoV-19/USA/LA-EVTL1216/2020\|EPI_ISL_768485\|2020-1 | .................... | .................... | .................... | .................... | .................... | .................... |
| AB | 3456 hCoV-19/USA/LA-EVTL1217/2020\|EPI_ISL_768486\|2020-1 | .................... | .................... | .................... | .................... | .................... | .................... |
| AB | 3457 hCoV-19/USA/LA-EVTL1218/2020\|EPI_ISL_768487\|2020-1 | .................... | .................... | .................... | .................... | .................... | .................... |
| AB | 3458 hCoV-19/USA/LA-EVTL1219/2020\|EPI_ISL_768488\|2020-1 | .................... | .................... | .................... | .................... | .................... | .................... |
| AB | 3459 hCoV-19/USA/LA-EVTL1220/2020\|EPI_ISL_768489\|2020-1 | .................... | .................... | .................... | .................... | .................... | .................... |
| AB | 3460 hCoV-19/USA/LA-EVTL1221/2020\|EPI_ISL_768490\|2020-1 | .................... | .................... | .................... | .................... | .................... | .................... |
| AB | 3461 hCoV-19/USA/LA-EVTL1224/2020\|EPI_ISL_768493\|2020-1 | .................... | .................... | .................... | .................... | .................... | .................... |
| AB | 3462 hCoV-19/USA/LA-EVTL1225/2020\|EPI_ISL_768494\|2020-1 | .................... | .A.................. | .................... | .................... | .................... | .................... |
| AB | 3463 hCoV-19/USA/LA-EVTL1226/2020\|EPI_ISL_768495\|2020-1 | .................... | .A.................. | .................... | .................... | .................... | .................... |
| AB | 3464 hCoV-19/USA/LA-EVTL1227/2020\|EPI_ISL_768496\|2020-1 | .................... | .A.................. | .................... | .................... | .................... | .................... |
| AB | 3465 hCoV-19/USA/LA-EVTL1228/2020\|EPI_ISL_768497\|2020-1 | .................... | .................... | .................... | .................... | .................... | .................... |
| AB | 3466 hCoV-19/USA/LA-EVTL1229/2020\|EPI_ISL_768498\|2020-1 | .................... | .A.................. | .................... | .................... | .................... | .................... |
| AB | 3467 hCoV-19/USA/LA-EVTL1230/2020\|EPI_ISL_768499\|2020-1 | .................... | .................... | .................... | .................... | .................... | .................... |
| AB | 3468 hCoV-19/England/MILK-C4FEA5/2020\|EPI_ISL_769701\|20 | .................... | .................... | .................... | .................... | .................... | .................... |
| AB | 3469 hCoV-19/England/MILK-C5166C/2020\|EPI_ISL_769706\|20 | .................... | .................... | .................... | .................... | .................... | .................... |
| AB | 3470 hCoV-19/England/MILK-C51866/2020\|EPI_ISL_769707\|20 | .................... | .................... | .................... | .................... | .................... | .................... |
| AB | 3471 hCoV-19/England/MILK-C539AD/2020\|EPI_ISL_769709\|20 | .................... | .................... | .................... | .................... | .................... | .................... |
| AB | 3472 hCoV-19/England/MILK-C4FD20/2020\|EPI_ISL_769710\|20 | .................... | .................... | .................... | .................... | .................... | .................... |
| AB | 3473 hCoV-19/England/QEUH-C4B8F6/2020\|EPI_ISL_769711\|20 | .................... | .................... | ..A................. | .................... | .................... | .................... |
| AB | 3474 hCoV-19/England/QEUH-C4B999/2020\|EPI_ISL_769714\|20 | .................... | .................... | .................... | .................... | .....T.............. | .................... |
| AB | 3475 hCoV-19/England/QEUH-C4BA87/2020\|EPI_ISL_769751\|20 | .................... | .................... | .................... | .................... | .................... | .................... |
| AB | 3476 hCoV-19/England/QEUH-C4B753/2020\|EPI_ISL_769759\|20 | .................... | .................... | .................... | .................... | .................... | .................... |
| AB | 3477 hCoV-19/England/MILK-C53891/2020\|EPI_ISL_769760\|20 | .................... | .................... | .................... | .................... | .................... | .................... |
| AB | 3478 hCoV-19/England/MILK-C515D8/2020\|EPI_ISL_769763\|20 | .................... | .................... | .................... | .................... | .................... | .................... |
| AB | 3479 hCoV-19/England/MILK-C53442/2020\|EPI_ISL_769766\|20 | .................... | .................... | .................... | .................... | .................... | .................... |
| AB | 3480 hCoV-19/England/MILK-C4FDB7/2020\|EPI_ISL_769767\|20 | .................... | .................... | .................... | .................... | .................... | .................... |
| AB | 3481 hCoV-19/England/QEUH-C4B96C/2020\|EPI_ISL_769768\|20 | .................... | .................... | .................... | .................... | .................... | .................... |
| AB | 3482 hCoV-19/England/MILK-C5014E/2020\|EPI_ISL_769772\|20 | .................... | .................... | .................... | .................... | .................... | .................... |
| AB | 3483 hCoV-19/England/QEUH-C4B771/2020\|EPI_ISL_769773\|20 | .................... | .................... | .................... | .................... | .................... | .................... |
| AB | 3484 hCoV-19/England/MILK-C501A8/2020\|EPI_ISL_769775\|20 | .................... | .................... | .................... | .................... | .....T.............. | .................... |
| AB | 3485 hCoV-19/England/MILK-C4FDD5/2020\|EPI_ISL_769778\|20 | .................... | .................... | .................... | .................... | .................... | .................... |
| AB | 3486 hCoV-19/England/MILK-C5018A/2020\|EPI_ISL_769779\|20 | .................... | .................... | .................... | .................... | .................... | .................... |
| AB | 3487 hCoV-19/England/MILK-C51A06/2020\|EPI_ISL_769781\|20 | .................... | .................... | .................... | .................... | .................... | .................... |
| AB | 3488 hCoV-19/England/MILK-C4FF93/2020\|EPI_ISL_769784\|20 | .................... | .................... | .................... | .................... | .................... | .................... |
| AB | 3489 hCoV-19/England/MILK-C4FE1E/2020\|EPI_ISL_769786\|20 | .................... | .................... | .................... | .................... | .................... | .................... |
| AB | 3490 hCoV-19/England/QEUH-C4B832/2020\|EPI_ISL_769787\|20 | .................... | .................... | .................... | .................... | .................... | .................... |
| AB | 3491 hCoV-19/England/QEUH-C4B95D/2020\|EPI_ISL_769788\|20 | .................... | .................... | .................... | .................... | .................... | .................... |
| AB | 3492 hCoV-19/England/MILK-C5399E/2020\|EPI_ISL_769793\|20 | .................... | .A.................. | .................... | .................... | .................... | ..A................. |
| AB | 3493 hCoV-19/England/QEUH-C4B8BA/2020\|EPI_ISL_769794\|20 | .................... | .................... | .................... | .................... | .................... | .................... |
| AB | 3494 hCoV-19/England/MILK-C5016C/2020\|EPI_ISL_769799\|20 | .................... | .................... | .................... | .................... | .....T.............. | .................... |
| AB | 3495 hCoV-19/Germany/NW-HHU-506/2020\|EPI_ISL_770722\|202 | .................... | .....A.............. | .................... | .................... | .................... | .................... |
| AB | 3496 hCoV-19/Germany/NW-HHU-510/2020\|EPI_ISL_770726\|202 | .................... | .................... | .................... | .................... | .................... | .................... |
| AB | 3497 hCoV-19/Germany/NW-HHU-511/2020\|EPI_ISL_770727\|202 | .................... | .....A.............. | .................... | .................... | .................... | .................... |
| AB | 3498 hCoV-19/Germany/NW-HHU-512/2020\|EPI_ISL_770728\|202 | .................... | .................... | .................... | .................... | .................... | .................... |
| AB | 3499 hCoV-19/Germany/NW-HHU-492/2020\|EPI_ISL_770741\|202 | .................... | .................... | .................... | .................... | .................... | .................... |
| AB | 3500 hCoV-19/Germany/NW-HHU-494/2020\|EPI_ISL_770743\|202 | .................... | .................... | .................... | .................... | .................... | .................... |
| AB | 3501 hCoV-19/Germany/NW-HHU-496/2020\|EPI_ISL_770745\|202 | .................... | .................... | .................... | .................... | .................... | .................... |
| AB | 3502 hCoV-19/Germany/NW-HHU-497/2020\|EPI_ISL_770746\|202 | .................... | .................... | .................... | .................... | .................... | .................... |
| AB | 3503 hCoV-19/Germany/NW-HHU-499/2020\|EPI_ISL_770747\|202 | .................... | .................... | .................... | .................... | .................... | .................... |
| AB | 3504 hCoV-19/Germany/NW-HHU-500/2020\|EPI_ISL_770748\|202 | .................... | .................... | .................... | .................... | .................... | .................... |
| AB | 3505 hCoV-19/Germany/NW-HHU-503/2020\|EPI_ISL_770751\|202 | .................... | .................... | .................... | .................... | .................... | .................... |

| | | | | | | | |
|---|---|---|---|---|---|---|---|
| AB | 3506 hCoV-19/Germany/NW-HHU-277/2020\|EPI_ISL_770752\|202 | .................... | .................... | .................... | .................... | .................... | ...............A.... |
| AB | 3507 hCoV-19/Germany/NW-HHU-278/2020\|EPI_ISL_770753\|202 | .................... | .................... | .................... | .................... | .................... | .................... |
| AB | 3508 hCoV-19/Germany/NW-HHU-280/2020\|EPI_ISL_770755\|202 | .................... | .................... | .................... | .................... | .................... | .................... |
| AB | 3509 hCoV-19/USA/OH-ODH-SC35023/2020\|EPI_ISL_770775\|202 | .................... | .................... | .................... | .................... | .................... | .................... |
| AB | 3510 hCoV-19/USA/OH-ODH-SC35024/2020\|EPI_ISL_770776\|202 | .................... | .................... | .................... | .................... | .................... | .................... |
| AB | 3511 hCoV-19/USA/OH-ODH-SC35046/2020\|EPI_ISL_770777\|202 | .................... | .................... | .................... | .................... | .................... | .................... |
| AB | 3512 hCoV-19/USA/MN-MDH-2204/2020\|EPI_ISL_770778\|2020-1 | .................... | .................... | .................... | .................... | .................... | .................... |
| AB | 3513 hCoV-19/USA/MN-MDH-2205/2020\|EPI_ISL_770779\|2020-1 | .................... | .................... | .................... | .................... | .................... | .................... |
| AB | 3514 hCoV-19/USA/MN-MDH-2206/2020\|EPI_ISL_770780\|2020-1 | .................... | .................... | .................... | .................... | .................... | .................... |
| AB | 3515 hCoV-19/USA/MN-MDH-2207/2020\|EPI_ISL_770781\|2020-1 | .................... | .................... | .................... | .................... | .................... | .................... |
| AB | 3516 hCoV-19/USA/MN-MDH-2208/2020\|EPI_ISL_770782\|2020-1 | .................... | .................... | .................... | .................... | .................... | .................... |
| AB | 3517 hCoV-19/USA/MN-MDH-2209/2020\|EPI_ISL_770783\|2020-1 | .................... | .................... | .................... | .................... | .................... | .................... |
| AB | 3518 hCoV-19/USA/MN-MDH-2210/2020\|EPI_ISL_770784\|2020-1 | .................... | .................... | .................... | .................... | .................... | .................... |
| AB | 3519 hCoV-19/USA/MN-MDH-2211/2020\|EPI_ISL_770785\|2020-1 | .................... | .................... | .................... | .................... | .................... | .................... |
| AB | 3520 hCoV-19/USA/MN-MDH-2212/2020\|EPI_ISL_770786\|2020-1 | .................... | .................... | .................... | .................... | .................... | .................... |
| AB | 3521 hCoV-19/USA/MN-MDH-2213/2020\|EPI_ISL_770787\|2020-1 | .................... | .................... | .................... | .................... | .................... | .................... |
| AB | 3522 hCoV-19/USA/MN-MDH-2214/2020\|EPI_ISL_770788\|2020-1 | .................... | .................... | .................... | .................... | .................... | .................... |
| AB | 3523 hCoV-19/USA/MN-MDH-2215/2020\|EPI_ISL_770789\|2020-1 | .................... | .................... | .................... | .................... | .................... | .................... |
| AB | 3524 hCoV-19/USA/MN-MDH-2216/2020\|EPI_ISL_770790\|2020-1 | .................... | .................... | .................... | .................... | .................... | .................... |
| AB | 3525 hCoV-19/USA/MN-MDH-2217/2020\|EPI_ISL_770791\|2020-1 | .................... | .................... | .................... | .................... | .................... | .................... |
| AB | 3526 hCoV-19/USA/MN-MDH-2218/2020\|EPI_ISL_770792\|2020-1 | .................... | .................... | .................... | .................... | .................... | .................... |
| AB | 3527 hCoV-19/USA/MN-MDH-2219/2020\|EPI_ISL_770793\|2020-1 | .................... | .A.................. | .................... | .................... | .NN................. | .................... |
| AB | 3528 hCoV-19/USA/MN-MDH-2220/2020\|EPI_ISL_770794\|2020-1 | .................... | .A.................. | .................... | .................... | .................... | .................... |
| AB | 3529 hCoV-19/USA/MN-MDH-2221/2020\|EPI_ISL_770795\|2020-1 | .................... | .................... | .................... | .................... | .................... | .................... |
| AB | 3530 hCoV-19/USA/MN-MDH-2222/2020\|EPI_ISL_770796\|2020-1 | .................... | .................... | .................... | .................... | .................... | .................... |
| AB | 3531 hCoV-19/USA/MN-MDH-2223/2020\|EPI_ISL_770797\|2020-0 | .................... | .A.................. | .................... | .................... | .................... | .................... |
| AB | 3532 hCoV-19/USA/MN-MDH-2224/2020\|EPI_ISL_770798\|2020-1 | .................... | .................... | .................... | .................... | .................... | .................... |
| AB | 3533 hCoV-19/USA/MN-MDH-2225/2020\|EPI_ISL_770799\|2020-1 | .................... | .................... | .................... | .................... | .................... | .................... |
| AB | 3534 hCoV-19/USA/OH-ODH-SC35064/2020\|EPI_ISL_775601\|202 | .................... | .................... | .................... | .................... | .................... | .................... |
| AB | 3535 hCoV-19/USA/OH-ODH-SC35065/2020\|EPI_ISL_775602\|202 | .................... | .................... | .................... | .................... | .................... | .................... |
| AB | 3536 hCoV-19/USA/OH-ODH-SC35067/2020\|EPI_ISL_775603\|202 | .................... | .................... | .................... | .................... | .................... | .................... |
| AB | 3537 hCoV-19/USA/OH-ODH-SC35076/2020\|EPI_ISL_775605\|202 | .................... | .................... | .................... | .................... | .................... | .................... |
| AB | 3538 hCoV-19/USA/OH-ODH-SC35077/2020\|EPI_ISL_775606\|202 | .................... | .................... | .................... | .................... | .................... | .................... |
| AB | 3539 hCoV-19/Italy/CAM-TIGEM-295/2020\|EPI_ISL_776925\|20 | .................... | .................... | .................... | .................... | .....T.............. | .................... |
| AB | 3540 hCoV-19/Italy/CAM-TIGEM-292/2020\|EPI_ISL_776926\|20 | .................... | .................... | .................... | .................... | .................... | .................... |
| AB | 3541 hCoV-19/Italy/CAM-TIGEM-252/2020\|EPI_ISL_776928\|20 | .................... | .................... | .................... | .................... | .................... | .................... |
| AB | 3542 hCoV-19/Italy/CAM-TIGEM-284/2020\|EPI_ISL_776932\|20 | .................... | .A.................. | .................... | .................... | .................... | .................... |
| AB | 3543 hCoV-19/Italy/CAM-TIGEM-293/2020\|EPI_ISL_776933\|20 | .................... | .................... | .................... | .................... | .................... | .................... |
| AB | 3544 hCoV-19/Italy/CAM-TIGEM-281/2020\|EPI_ISL_776934\|20 | .................... | .................... | .................... | .................... | .....T.............. | .................... |
| AB | 3545 hCoV-19/Italy/CAM-TIGEM-286/2020\|EPI_ISL_776935\|20 | .................... | .................... | .................... | .................... | .................... | .................... |
| AB | 3546 hCoV-19/Italy/CAM-TIGEM-253/2020\|EPI_ISL_776936\|20 | .................... | .................... | .................... | .................... | .................... | .................... |
| AB | 3547 hCoV-19/Italy/CAM-TIGEM-279/2020\|EPI_ISL_776937\|20 | .................... | .................... | .................... | .................... | .................... | .................... |
| AB | 3548 hCoV-19/Italy/CAM-TIGEM-258/2020\|EPI_ISL_776938\|20 | .................... | .................... | .................... | .................... | .................... | .................... |
| AB | 3549 hCoV-19/Italy/CAM-TIGEM-294/2020\|EPI_ISL_776939\|20 | .................... | .A.................. | .................... | .................... | .................... | .................... |
| AB | 3550 hCoV-19/Italy/CAM-TIGEM-259/2020\|EPI_ISL_776940\|20 | .................... | .................... | .................... | .................... | .................... | .................... |
| AB | 3551 hCoV-19/Italy/CAM-TIGEM-255/2020\|EPI_ISL_776941\|20 | .................... | .................... | .................... | .................... | .....T.............. | .................... |
| AB | 3552 hCoV-19/Italy/CAM-TIGEM-278/2020\|EPI_ISL_776942\|20 | .................... | .................... | .................... | .................... | .................... | .................... |
| AB | 3553 hCoV-19/USA/LA-EVTL1434/2020\|EPI_ISL_779100\|2020-1 | .................... | .................... | .................... | .................... | .................... | .................... |
| AB | 3554 hCoV-19/USA/LA-EVTL1435/2020\|EPI_ISL_779101\|2020-1 | .................... | .................... | .................... | .................... | .................... | .................... |
| AB | 3555 hCoV-19/USA/LA-EVTL1436/2020\|EPI_ISL_779102\|2020-1 | .................... | .................... | .................... | .................... | .................... | .................... |
| AB | 3556 hCoV-19/USA/LA-EVTL1437/2020\|EPI_ISL_779103\|2020-1 | .................... | .................... | .................... | .................... | .................... | .................... |
| AB | 3557 hCoV-19/USA/LA-EVTL1438/2020\|EPI_ISL_779104\|2020-1 | .................... | .................... | .................... | .................... | .................... | .................... |
| AB | 3558 hCoV-19/USA/LA-EVTL1439/2020\|EPI_ISL_779105\|2020-1 | .................... | .................... | .................... | .................... | .................... | .................... |
| AB | 3559 hCoV-19/USA/LA-EVTL1440/2020\|EPI_ISL_779106\|2020-1 | .................... | .................... | .................... | .................... | .................... | .................... |
| AB | 3560 hCoV-19/USA/LA-EVTL1441/2020\|EPI_ISL_779107\|2020-1 | .................... | .................... | .................... | .................... | .................... | .................... |
| AB | 3561 hCoV-19/USA/LA-EVTL1442/2020\|EPI_ISL_779108\|2020-1 | .................... | .................... | .................... | .................... | .................... | .................... |
| AB | 3562 hCoV-19/USA/LA-EVTL1443/2020\|EPI_ISL_779109\|2020-1 | .................... | .................... | .................... | .................... | .................... | .................... |
| AB | 3563 hCoV-19/USA/LA-EVTL1444/2020\|EPI_ISL_779110\|2020-1 | .................... | .................... | .................... | .................... | .................... | .................... |
| AB | 3564 hCoV-19/USA/LA-EVTL1445/2020\|EPI_ISL_779111\|2020-1 | .................... | .................... | .................... | .................... | .................... | .................... |
| AB | 3565 hCoV-19/USA/LA-EVTL1446/2020\|EPI_ISL_779112\|2020-1 | .................... | .................... | .................... | .................... | .................... | .................... |
| AB | 3566 hCoV-19/USA/LA-EVTL1447/2020\|EPI_ISL_779113\|2020-1 | .................... | .A.................. | .................... | .................... | .................... | .................... |
| AB | 3567 hCoV-19/USA/LA-EVTL1448/2020\|EPI_ISL_779114\|2020-1 | .................... | .................... | .................... | .................... | .................... | .................... |
| AB | 3568 hCoV-19/USA/LA-EVTL1449/2020\|EPI_ISL_779115\|2020-1 | .................... | .................... | .................... | .................... | .................... | .................... |
| AB | 3569 hCoV-19/USA/LA-EVTL1450/2020\|EPI_ISL_779116\|2020-1 | .................... | .................... | .................... | .................... | .................... | .................... |
| AB | 3570 hCoV-19/USA/LA-EVTL1451/2020\|EPI_ISL_779117\|2020-1 | .................... | .................... | .................... | .................... | .................... | .................... |
| AB | 3571 hCoV-19/USA/LA-EVTL1452/2020\|EPI_ISL_779118\|2020-1 | .................... | .................... | .................... | .................... | .................... | .................... |
| AB | 3572 hCoV-19/USA/LA-EVTL1453/2020\|EPI_ISL_779119\|2020-1 | .................... | .................... | .................... | .................... | .................... | .................... |
| AB | 3573 hCoV-19/USA/LA-EVTL1454/2020\|EPI_ISL_779120\|2020-1 | .................... | .A.................. | .................... | .................... | .................... | .................... |
| AB | 3574 hCoV-19/USA/LA-EVTL1456/2020\|EPI_ISL_779122\|2020-1 | .................... | .A.................. | .................... | .................... | .................... | .................... |
| AB | 3575 hCoV-19/USA/LA-EVTL1457/2020\|EPI_ISL_779123\|2020-1 | .................... | .A.................. | .................... | .................... | .................... | .................... |
| AB | 3576 hCoV-19/USA/LA-EVTL1458/2020\|EPI_ISL_779124\|2020-1 | .................... | .................... | .................... | .................... | .................... | .................... |
| AB | 3577 hCoV-19/USA/LA-EVTL1459/2020\|EPI_ISL_779125\|2020-1 | .................... | .................... | .................... | .................... | .................... | .................... |
| AB | 3578 hCoV-19/USA/LA-EVTL1460/2020\|EPI_ISL_779126\|2020-1 | .................... | .A.................. | .................... | .................... | .................... | .................... |
| AB | 3579 hCoV-19/USA/LA-EVTL1462/2020\|EPI_ISL_779128\|2020-1 | .................... | .................... | .................... | .................... | .................... | .................... |
| AB | 3580 hCoV-19/Brazil/RS-00601/2020\|EPI_ISL_779155\|2020-1 | .................... | .................... | .................... | .................... | .................... | .................... |
| AB | 3581 hCoV-19/Brazil/RS-00602/2020\|EPI_ISL_779156\|2020-1 | .................... | .....A.............. | .................... | .................... | .................... | .................... |
| AB | 3582 hCoV-19/Brazil/RS-00621/2020\|EPI_ISL_779157\|2020-1 | .................... | .................... | .................... | .................... | .................... | .................... |
| AB | 3583 hCoV-19/Brazil/RS-00624/2020\|EPI_ISL_779158\|2020-1 | .................... | .................... | .................... | .................... | .................... | .................... |
| AB | 3584 hCoV-19/Brazil/RS-00631/2020\|EPI_ISL_779159\|2020-1 | .................... | .................... | .................... | .................... | .................... | .................... |
| AB | 3585 hCoV-19/England/ALDP-CAFA54/2020\|EPI_ISL_761001\|20 | .................... | .................... | .................... | .................... | .................... | .................... |

| | | | | | | | | |
|---|---|---|---|---|---|---|---|---|
| AB | 3586 hCoV-19/England/MILK-C201ED/2020\|EPI_ISL_761002\|20 | ···················· | ···················· | ···················· | ···················· | ···················· | ···················· | |
| AB | 3587 hCoV-19/England/QEUH-CAD0DC/2020\|EPI_ISL_761005\|20 | ···················· | ···················· | ···················· | ···················· | ···················· | ···················· | |
| AB | 3588 hCoV-19/England/QEUH-CADCE7/2020\|EPI_ISL_761007\|20 | ···················· | ···················· | ···················· | ···················· | ······T············· | ···················· | |
| AB | 3589 hCoV-19/England/QEUH-CAD1CA/2020\|EPI_ISL_761010\|20 | ···················· | ·········T·········· | ···················· | ···················· | ···················· | ···················· | |
| AB | 3590 hCoV-19/England/ALDP-CB456A/2020\|EPI_ISL_761011\|20 | ···················· | ···················· | ···················· | ···················· | ···················· | ···················· | |
| AB | 3591 hCoV-19/England/QEUH-CADCF6/2020\|EPI_ISL_761012\|20 | ···················· | ···················· | ···················· | ···················· | ···················· | ···················· | |
| AB | 3592 hCoV-19/England/ALDP-CB2F6E/2020\|EPI_ISL_761013\|20 | ···················· | ···················· | ···················· | ···················· | ···················· | ···················· | |
| AB | 3593 hCoV-19/England/QEUH-CADA29/2020\|EPI_ISL_761014\|20 | ···················· | ···················· | ···················· | ···················· | ···················· | ···················· | |
| AB | 3594 hCoV-19/England/QEUH-CAD8D4/2020\|EPI_ISL_761015\|20 | ···················· | ···················· | ···················· | ···················· | ···················· | ···················· | |
| AB | 3595 hCoV-19/England/CAMC-C12B2C/2020\|EPI_ISL_761017\|20 | ···················· | ···················· | ···················· | ···················· | ···················· | ···················· | |
| AB | 3596 hCoV-19/England/QEUH-C4CC35/2020\|EPI_ISL_761018\|20 | ···················· | ···················· | ···················· | ···················· | ···················· | ···················· | |
| AB | 3597 hCoV-19/England/CAMC-BF84E6/2020\|EPI_ISL_761021\|20 | ···················· | ···················· | ···················· | ···················· | ···················· | ···················· | |
| AB | 3598 hCoV-19/England/ALDP-CB42CD/2020\|EPI_ISL_761022\|20 | ···················· | ···················· | ···················· | ···················· | ···················· | ···················· | |
| AB | 3599 hCoV-19/England/MILK-CB28EB/2020\|EPI_ISL_761023\|20 | ···················· | ···················· | ···················· | ···················· | ···················· | ···················· | |
| AB | 3600 hCoV-19/England/QEUH-CADA0B/2020\|EPI_ISL_761026\|20 | ···················· | ···················· | ···················· | ···················· | ···················· | ···················· | |
| AB | 3601 hCoV-19/England/QEUH-CAD4A3/2020\|EPI_ISL_761030\|20 | ···················· | ···················· | ···················· | ···················· | ···················· | ···················· | |
| AB | 3602 hCoV-19/England/QEUH-CADE69/2020\|EPI_ISL_761031\|20 | ··················T· | ···················· | ···················· | ···················· | ···················· | ···················· | 1 |
| AB | 3603 hCoV-19/England/QEUH-CADE78/2020\|EPI_ISL_761035\|20 | ···················· | ···················· | ···················· | ···················· | ···················· | ···················· | |
| AB | 3604 hCoV-19/England/MILK-CB2809/2020\|EPI_ISL_761039\|20 | ···················· | ···················· | ···················· | ···················· | ···················· | ···················· | |
| AB | 3605 hCoV-19/England/ALDP-CB4E23/2020\|EPI_ISL_761040\|20 | ···················· | ···················· | ···················· | ···················· | ···················· | ···················· | |
| AB | 3606 hCoV-19/England/ALDP-CB2E16/2020\|EPI_ISL_761042\|20 | ···················· | ···················· | ···················· | ···················· | ···················· | ···················· | |
| AB | 3607 hCoV-19/England/QEUH-CB31D0/2020\|EPI_ISL_761043\|20 | ···················· | ···················· | ···················· | ···················· | ···················· | ···················· | |
| AB | 3608 hCoV-19/England/QEUH-CAD106/2020\|EPI_ISL_761045\|20 | ···················· | ···················· | ···················· | ···················· | ···················· | ···················· | |
| AB | 3609 hCoV-19/England/ALDP-CB3228/2020\|EPI_ISL_761046\|20 | ···················· | ···················· | ···················· | ···················· | ···················· | ···················· | |
| AB | 3610 hCoV-19/England/QEUH-CB0F51/2020\|EPI_ISL_761047\|20 | ···················· | ···················· | ···················· | ···················· | ···················· | ···················· | |
| AB | 3611 hCoV-19/England/ALDP-CB4236/2020\|EPI_ISL_761049\|20 | ···················· | ···················· | ···················· | ···················· | ···················· | ···················· | |
| AB | 3612 hCoV-19/England/MILK-CB23CC/2020\|EPI_ISL_761050\|20 | ···················· | ···················· | ···················· | ···················· | ···················· | ···················· | |
| AB | 3613 hCoV-19/England/QEUH-CAD86B/2020\|EPI_ISL_761051\|20 | ···················· | ···················· | ···················· | ···················· | ···················· | ···················· | |
| AB | 3614 hCoV-19/England/QEUH-CAD9FF/2020\|EPI_ISL_761053\|20 | ···················· | ···················· | ···················· | ···················· | ···················· | ···················· | |
| AB | 3615 hCoV-19/England/ALDP-CB45A6/2020\|EPI_ISL_761054\|20 | ···················· | ···················· | ···················· | ···················· | ···················· | ···················· | |
| AB | 3616 hCoV-19/England/QEUH-CABDE6/2020\|EPI_ISL_761058\|20 | ···················· | ···················· | ···················· | ···················· | ······T············· | ···················· | |
| AB | 3617 hCoV-19/England/ALDP-CB42BE/2020\|EPI_ISL_761060\|20 | ···················· | ···················· | ···················· | ···················· | ···A················ | ···················· | |
| AB | 3618 hCoV-19/England/ALDP-C05342/2020\|EPI_ISL_761061\|20 | ···················· | ···················· | ···················· | ···················· | ···················· | ···················· | |
| AB | 3619 hCoV-19/England/QEUH-CABE10/2020\|EPI_ISL_761063\|20 | ···················· | ····A··············· | ···················· | ···················· | ···················· | ···················· | |
| AB | 3620 hCoV-19/England/ALDP-CB3F21/2020\|EPI_ISL_761064\|20 | ···················· | ···················· | ···················· | ···················· | ···················· | ···················· | |
| AB | 3621 hCoV-19/England/ALDP-CB46EF/2020\|EPI_ISL_761065\|20 | ···················· | ···················· | ···················· | ···················· | ···················· | ···················· | |
| AB | 3622 hCoV-19/England/ALDP-CB3D63/2020\|EPI_ISL_761066\|20 | ···················· | ···················· | ···················· | ···················· | ···················· | ···················· | |
| AB | 3623 hCoV-19/England/QEUH-CB338F/2020\|EPI_ISL_761067\|20 | ···················· | ·A·················· | ···················· | ···················· | ···················· | ···A················ | |
| AB | 3624 hCoV-19/England/QEUH-CADA83/2020\|EPI_ISL_761068\|20 | ···················· | ···················· | ···················· | ···················· | ···················· | ···················· | |
| AB | 3625 hCoV-19/England/QEUH-CAB539/2020\|EPI_ISL_761072\|20 | ···················· | ···················· | ···················· | ···················· | ···················· | ···················· | |
| AB | 3626 hCoV-19/England/QEUH-CAD221/2020\|EPI_ISL_761073\|20 | ···················· | ···················· | ···················· | ···················· | ···················· | ···················· | |
| AB | 3627 hCoV-19/England/CAMC-C1E8A1/2020\|EPI_ISL_761074\|20 | ···················· | ···················· | ···················· | ···················· | ···················· | ···················· | |
| AB | 3628 hCoV-19/England/CAMC-C131CF/2020\|EPI_ISL_761075\|20 | ···················· | ·A·················· | ···················· | ···················· | ···················· | ···A················ | |
| AB | 3629 hCoV-19/England/QEUH-CAD9D1/2020\|EPI_ISL_761076\|20 | ···················· | ·A·················· | ···················· | ···················· | ···················· | ···················· | |
| AB | 3630 hCoV-19/England/QEUH-CABE2F/2020\|EPI_ISL_761078\|20 | ···················· | ···················· | ···················· | ···················· | ···················· | ···················· | |
| AB | 3631 hCoV-19/England/MILK-CB0364/2020\|EPI_ISL_761079\|20 | ···················· | ···················· | ···················· | ···················· | ···················· | ···················· | |
| AB | 3632 hCoV-19/England/MILK-CA9A87/2020\|EPI_ISL_762301\|20 | ···················· | ···················· | ···················· | ···················· | ···················· | ···················· | |
| AB | 3633 hCoV-19/England/MILK-CA9C81/2020\|EPI_ISL_762311\|20 | ···················· | ···················· | ···················· | ···················· | ···················· | ···················· | |
| AB | 3634 hCoV-19/England/MILK-CA459A/2020\|EPI_ISL_762318\|20 | ···T················ | ···················· | ···················· | ···················· | ···················· | ···················· | |
| AB | 3635 hCoV-19/England/MILK-CA960B/2020\|EPI_ISL_762322\|20 | ···················· | ···················· | ···················· | ···················· | ···················· | ···················· | |
| AB | 3636 hCoV-19/England/MILK-CA9902/2020\|EPI_ISL_762323\|20 | ···················· | ···················· | ···················· | ···················· | ···················· | ···················· | |
| AB | 3637 hCoV-19/England/MILK-CA4B2F/2020\|EPI_ISL_762324\|20 | ···················· | ···················· | ···················· | ···················· | ···················· | ···················· | |
| AB | 3638 hCoV-19/England/MILK-CA41A5/2020\|EPI_ISL_762326\|20 | ···················· | ···················· | ···················· | ···················· | ···················· | ···················· | |
| AB | 3639 hCoV-19/England/MILK-CA513B/2020\|EPI_ISL_762327\|20 | ···················· | ···················· | ···················· | ···················· | ···················· | ···················· | |
| AB | 3640 hCoV-19/England/MILK-CA9D15/2020\|EPI_ISL_762329\|20 | ···················· | ···················· | ···················· | ···················· | ···················· | ···················· | |
| AB | 3641 hCoV-19/England/MILK-CA9CEB/2020\|EPI_ISL_762335\|20 | ···················· | ···················· | ···················· | ···················· | ······T············· | ···················· | |
| AB | 3642 hCoV-19/England/MILK-CA9823/2020\|EPI_ISL_762341\|20 | ···················· | ···················· | ···················· | ···················· | ······T············· | ···················· | |
| AB | 3643 hCoV-19/England/MILK-CA9B1B/2020\|EPI_ISL_762354\|20 | ···················· | ···················· | ···················· | ···················· | ···················· | ···················· | |
| AB | 3644 hCoV-19/England/MILK-CA9D9D/2020\|EPI_ISL_762374\|20 | ···················· | ···················· | ···················· | ···················· | ···················· | ···················· | |
| AB | 3645 hCoV-19/England/MILK-CA4196/2020\|EPI_ISL_762389\|20 | ···················· | ···················· | ···················· | ···················· | ···················· | ···················· | |
| AB | 3646 hCoV-19/England/MILK-CA3E54/2020\|EPI_ISL_762398\|20 | ···················· | ···················· | ···················· | ···················· | ···················· | ···················· | |
| AB | 3647 hCoV-19/England/MILK-CA371D/2020\|EPI_ISL_762399\|20 | ···················· | ···················· | ···················· | ···················· | ···················· | ···················· | |
| AB | 3648 hCoV-19/Wales/PHWC-4A5670/2020\|EPI_ISL_764900\|2020 | ···················· | ···················· | ···················· | ···················· | ···················· | ···················· | |
| AB | 3649 hCoV-19/Wales/PHWC-4A568F/2020\|EPI_ISL_764901\|2020 | ···················· | ···················· | ···················· | ···················· | ···················· | ···················· | |
| AB | 3650 hCoV-19/Wales/PHWC-4A569E/2020\|EPI_ISL_764902\|2020 | ···················· | ···················· | ···················· | ···················· | ···················· | ···················· | |
| AB | 3651 hCoV-19/Wales/PHWC-4A56AD/2020\|EPI_ISL_764903\|2020 | ···················· | ···················· | ···················· | ···················· | ···················· | ···················· | |
| AB | 3652 hCoV-19/Wales/PHWC-4A56BC/2020\|EPI_ISL_764904\|2020 | ···················· | ···················· | ···················· | ···················· | ···················· | ···················· | |
| AB | 3653 hCoV-19/Wales/PHWC-4A56CB/2020\|EPI_ISL_764905\|2020 | ···················· | ···················· | ···················· | ···················· | ···················· | ···················· | |
| AB | 3654 hCoV-19/Wales/PHWC-4A56DA/2020\|EPI_ISL_764906\|2020 | ···················· | ···················· | ···················· | ···················· | ···················· | ···················· | |
| AB | 3655 hCoV-19/Wales/PHWC-4A56E9/2020\|EPI_ISL_764907\|2020 | ···················· | ···················· | ···················· | ···················· | ···················· | ···················· | |
| AB | 3656 hCoV-19/Wales/PHWC-4A56F8/2020\|EPI_ISL_764908\|2020 | ···················· | ···················· | ···················· | ···················· | ···················· | ···················· | |
| AB | 3657 hCoV-19/Wales/PHWC-4A5704/2020\|EPI_ISL_764909\|2020 | ···················· | ···················· | ···················· | ···················· | ···················· | ···················· | |
| AB | 3658 hCoV-19/Wales/PHWC-4A5713/2020\|EPI_ISL_764910\|2020 | ···················· | ···················· | ···················· | ···················· | ···················· | ···················· | |
| AB | 3659 hCoV-19/Wales/PHWC-4A5722/2020\|EPI_ISL_764911\|2020 | ···················· | ···················· | ···················· | ···················· | ···················· | ···················· | |
| AB | 3660 hCoV-19/Wales/PHWC-4A5731/2020\|EPI_ISL_764912\|2020 | ···················· | ···················· | ···················· | ···················· | ···················· | ···················· | |
| AB | 3661 hCoV-19/Wales/PHWC-4A5740/2020\|EPI_ISL_764913\|2020 | ···················· | ···················· | ···················· | ···················· | ···················· | ···················· | |
| AB | 3662 hCoV-19/Wales/PHWC-4A575F/2020\|EPI_ISL_764914\|2020 | ···················· | ···················· | ···················· | ···················· | ···················· | ···················· | |
| AB | 3663 hCoV-19/Wales/PHWC-4A576E/2020\|EPI_ISL_764915\|2020 | ···················· | ···················· | ···················· | ···················· | ···················· | ···················· | |
| AB | 3664 hCoV-19/Wales/PHWC-4A577D/2020\|EPI_ISL_764916\|2020 | ···················· | ···················· | ···················· | ···················· | ···················· | ···················· | |
| AB | 3665 hCoV-19/Wales/PHWC-4A578C/2020\|EPI_ISL_764917\|2020 | ···················· | ···················· | ···················· | ···················· | ···················· | ···················· | |

AB 3666 hCoV-19/Wales/PHWC-4A579B/2020|EPI_ISL_764918|2020 ···················· ···················· ···················· ···················· ···················· ····················
AB 3667 hCoV-19/Wales/PHWC-4A57AA/2020|EPI_ISL_764919|2020 ···················· ···················· ···················· ···················· ···················· ····················
AB 3668 hCoV-19/Wales/PHWC-4A57B9/2020|EPI_ISL_764920|2020 ···················· ···················· ···················· ···················· ···T················ ····················
AB 3669 hCoV-19/Wales/PHWC-4A57C8/2020|EPI_ISL_764921|2020 ···················· ···················· ···················· ···················· ···················· ····················
AB 3670 hCoV-19/Wales/PHWC-4A57D7/2020|EPI_ISL_764922|2020 ···················· ···················· ···················· ···················· ···················· ····················
AB 3671 hCoV-19/Wales/PHWC-4A57E6/2020|EPI_ISL_764923|2020 ···················· ···················· ···················· ···················· ···················· ····················
AB 3673 hCoV-19/Wales/PHWC-4A5801/2020|EPI_ISL_764925|2020 ···················· ···················· ···················· ···················· ···················· ····················
AB 3674 hCoV-19/Wales/PHWC-4A582F/2020|EPI_ISL_764926|2020 ···················· ···················· ···················· ···················· ···················· ····················
AB 3675 hCoV-19/Wales/PHWC-4A583E/2020|EPI_ISL_764927|2020 ···················· ···················· ···················· ···················· ···················· ····················
AB 3676 hCoV-19/Wales/PHWC-4A584D/2020|EPI_ISL_764928|2020 ···················· ···················· ···················· ···················· ···················· ····················
AB 3677 hCoV-19/Wales/PHWC-4A585C/2020|EPI_ISL_764929|2020 ···················· ···················· ···················· ···················· ···················· ····················
AB 3678 hCoV-19/Wales/PHWC-4A586B/2020|EPI_ISL_764930|2020 ···················· ···················· ···················· ···················· ···················· ····················
AB 3679 hCoV-19/Wales/PHWC-4A587A/2020|EPI_ISL_764931|2020 ···················· ···················· ···················· ···················· ···················· ····················
AB 3680 hCoV-19/Wales/PHWC-4A5889/2020|EPI_ISL_764932|2020 ···················· ···················· ···················· ···················· ···················· ····················
AB 3681 hCoV-19/Wales/PHWC-4A5898/2020|EPI_ISL_764933|2020 ···················· ···················· ···················· ···················· ···················· ····················
AB 3682 hCoV-19/Wales/PHWC-4A58B6/2020|EPI_ISL_764934|2020 ···················· ···················· ···················· ···················· ···················· ····················
AB 3683 hCoV-19/Wales/PHWC-4A58C5/2020|EPI_ISL_764935|2020 ···················· ···················· ···················· ···················· ···················· ····················
AB 3684 hCoV-19/Wales/PHWC-4A58D4/2020|EPI_ISL_764936|2020 ···················· ···················· ···················· ···················· ···················· ····················
AB 3685 hCoV-19/Wales/PHWC-4A58E3/2020|EPI_ISL_764937|2020 ···················· ···················· ···················· ···················· ···················· ····················
AB 3692 hCoV-19/Wales/PHWC-4A5B44/2020|EPI_ISL_764955|2020 ···················· ···················· ···················· ···················· ···················· ····················
AB 3693 hCoV-19/Wales/PHWC-4A5B71/2020|EPI_ISL_764956|2020 ···················· ···················· ···················· ···················· ···················· ····················
AB 3695 hCoV-19/Wales/PHWC-4A5BBD/2020|EPI_ISL_764958|2020 ···················· ···················· ···················· ···················· ···················· ····················
AB 3697 hCoV-19/Wales/PHWC-4A5C14/2020|EPI_ISL_764960|2020 ···················· ···················· ···················· ···················· ···················· ····················
AB 3700 hCoV-19/Wales/PHWC-4A5C50/2020|EPI_ISL_764963|2020 ···················· ···················· ···················· ···················· ···················· ····················
AB 3702 hCoV-19/Wales/PHWC-4A5C8D/2020|EPI_ISL_764965|2020 ···················· ···················· ···················· ···················· ···················· ····················
AB 3704 hCoV-19/Wales/PHWC-4A5CF6/2020|EPI_ISL_764967|2020 ···················· ···················· ···················· ···················· ···················· ····················
AB 3707 hCoV-19/Wales/PHWC-4A5D20/2020|EPI_ISL_764970|2020 ···················· ···················· ···················· ···················· ···················· ····················
AB 3709 hCoV-19/Wales/PHWC-4A5D4E/2020|EPI_ISL_764972|2020 ···················· ···················· ···················· ···················· ···················· ····················
AB 3710 hCoV-19/Wales/PHWC-4A5D7B/2020|EPI_ISL_764973|2020 ···················· ···················· ···················· ···················· ···················· ····················
AB 3713 hCoV-19/Wales/PHWC-4A5E2D/2020|EPI_ISL_764976|2020 ···················· ···················· ···················· ···················· ···················· ····················
AB 3715 hCoV-19/Wales/PHWC-4A5E4B/2020|EPI_ISL_764978|2020 ···················· ···················· ···················· ···················· ···················· ····················
AB 3719 hCoV-19/Wales/PHWC-4A5F0C/2020|EPI_ISL_764982|2020 ···················· ···················· ···················· ···················· ···················· ····················
AB 3720 hCoV-19/Wales/PHWC-4A5F1B/2020|EPI_ISL_764983|2020 ···················· ···················· ···················· ···················· ···················· ····················
AB 3722 hCoV-19/Wales/PHWC-4A5F66/2020|EPI_ISL_764985|2020 ···················· ···················· ···················· ···················· ···················· ····················
AB 3723 hCoV-19/Wales/PHWC-4A5F75/2020|EPI_ISL_764986|2020 ···················· ···················· ···················· ···················· ···················· ····················
AB 3724 hCoV-19/Wales/PHWC-4A5F93/2020|EPI_ISL_764987|2020 ···················· ···················· ···················· ···················· ···················· ····················
AB 3725 hCoV-19/Wales/PHWC-4A5FA2/2020|EPI_ISL_764988|2020 ···················· ···················· ···················· ···················· ···················· ····················
AB 3726 hCoV-19/Wales/PHWC-4A5FDF/2020|EPI_ISL_764989|2020 ···················· ···················· ···················· ···················· ···················· ····················
AB 3728 hCoV-19/Wales/PHWC-4A6026/2020|EPI_ISL_764991|2020 ···················· ···················· ···················· ···················· ···················· ····················
AB 3729 hCoV-19/Wales/PHWC-4A6044/2020|EPI_ISL_764992|2020 ···················· ···················· ···················· ···················· ···················· ····················
AB 3730 hCoV-19/Wales/PHWC-4A6053/2020|EPI_ISL_764993|2020 ···················· ···················· ···················· ···················· ···················· ····················
AB 3732 hCoV-19/Wales/PHWC-4A6071/2020|EPI_ISL_764995|2020 ···················· ···················· ···················· ···················· ···················· ····················
AB 3733 hCoV-19/Wales/PHWC-4A60AE/2020|EPI_ISL_764996|2020 ···················· ···················· ···················· ···················· ···················· ····················
AB 3737 hCoV-19/Wales/ALDP-C39A13/2020|EPI_ISL_767180|2020 ···················· ···················· ···················· ···················· ···················· ····················
AB 3738 hCoV-19/Wales/ALDP-C3A520/2020|EPI_ISL_767181|2020 ···················· ···················· ···················· ···················· ···················· ····················
AB 3739 hCoV-19/Wales/ALDP-C3972B/2020|EPI_ISL_767182|2020 ···················· ···················· ···················· ···················· ···················· ····················
AB 3740 hCoV-19/Wales/ALDP-C399CB/2020|EPI_ISL_767183|2020 ···················· ···················· ···················· ···················· ···················· ····················
AB 3741 hCoV-19/Wales/ALDP-C399AD/2020|EPI_ISL_767184|2020 ···················· ···················· ···················· ···················· ···················· ····················
AB 3742 hCoV-19/Wales/ALDP-C3B361/2020|EPI_ISL_767185|2020 ···················· ···················· ···················· ···················· ···················· ····················
AB 3743 hCoV-19/Wales/ALDP-C3ADFB/2020|EPI_ISL_767186|2020 ···················· ···················· ···················· ···················· ···················· ····················
AB 3744 hCoV-19/Wales/ALDP-C3B46E/2020|EPI_ISL_767187|2020 ···················· ···················· ···················· ···················· ···················· ····················
AB 3745 hCoV-19/Wales/ALDP-C399E9/2020|EPI_ISL_767188|2020 ···················· ···················· ···················· ···················· ···················· ····················
AB 3746 hCoV-19/Wales/ALDP-C396E2/2020|EPI_ISL_767189|2020 ···················· ···················· ···················· ···················· ···················· ····················
AB 3747 hCoV-19/Wales/ALDP-C3970D/2020|EPI_ISL_767190|2020 ···················· ···················· ···················· ···················· ···················· ····················
AB 3748 hCoV-19/Wales/ALDP-C3A502/2020|EPI_ISL_767191|2020 ···················· ···················· ···················· ···················· ···················· ····················
AB 3749 hCoV-19/Wales/ALDP-C39952/2020|EPI_ISL_767192|2020 ···················· ···················· ···················· ···················· ···················· ····················
AB 3750 hCoV-19/Wales/ALDP-C39776/2020|EPI_ISL_767193|2020 ···················· ···················· ···················· ···················· ···················· ····················
AB 3751 hCoV-19/Wales/ALDP-C3971C/2020|EPI_ISL_767194|2020 ···················· ···················· ···················· ···················· ···················· ····················
AB 3752 hCoV-19/Wales/ALDP-C39A31/2020|EPI_ISL_767195|2020 ···················· ···················· ···················· ···················· ···················· ····················
AB 3753 hCoV-19/Wales/ALDP-C39970/2020|EPI_ISL_767196|2020 ···················· ···················· ···················· ···················· ···················· ····················
AB 3754 hCoV-19/Wales/ALDP-C39785/2020|EPI_ISL_767197|2020 ···················· ···················· ···················· ···················· ···················· ····················
AB 3755 hCoV-19/Wales/ALDP-C39A40/2020|EPI_ISL_767198|2020 ···················· ···················· ···················· ···················· ···················· ····················
AB 3756 hCoV-19/Wales/ALDP-C399BC/2020|EPI_ISL_767199|2020 ···················· ···················· ···················· ···················· ···················· ····················
AB 3757 hCoV-19/USA/OH-ODH-SC35063/2020|EPI_ISL_775600|202 ···················· ···················· ···················· ···················· ···················· ····················
AB 3758 hCoV-19/Italy/CAM-TIGEM-245/2020|EPI_ISL_776900|20 ···················· ·A·················· ···················· ···················· ···················· ····················
AB 3759 hCoV-19/Italy/CAM-TIGEM-250/2020|EPI_ISL_776901|20 ···················· ···················· ···················· ···················· ···················· ····················
AB 3760 hCoV-19/Italy/CAM-TIGEM-238/2020|EPI_ISL_776902|20 ···················· ···················· ···················· ···················· ···················· ····················
AB 3761 hCoV-19/Italy/CAM-TIGEM-226/2020|EPI_ISL_776903|20 ···················· ···················· ···················· ···················· ···················· ····················
AB 3762 hCoV-19/Italy/CAM-TIGEM-228/2020|EPI_ISL_776904|20 ···················· ·A·················· ···················· ···················· ···················· ····················
AB 3763 hCoV-19/Italy/CAM-TIGEM-242/2020|EPI_ISL_776905|20 ···················· ···················· ···················· ···················· ···················· ····················
AB 3764 hCoV-19/Italy/CAM-TIGEM-246/2020|EPI_ISL_776906|20 ···················· ···················· ···················· ···················· ···················· ····················
AB 3765 hCoV-19/Italy/CAM-TIGEM-232/2020|EPI_ISL_776907|20 ···················· ·A·················· ···················· ···················· ···················· ····················
AB 3766 hCoV-19/Italy/CAM-TIGEM-227/2020|EPI_ISL_776908|20 ···················· ·A·················· ···················· ···················· ···················· ····················
AB 3767 hCoV-19/Italy/CAM-TIGEM-224/2020|EPI_ISL_776909|20 ···················· ·A·················· ···················· ···················· ···················· ····················
AB 3768 hCoV-19/Italy/CAM-TIGEM-197/2020|EPI_ISL_776910|20 ···················· ···················· ···················· ···················· ···················· ····················
AB 3769 hCoV-19/Italy/CAM-TIGEM-237/2020|EPI_ISL_776911|20 ···················· ···················· ···················· ···················· ···················· ····················
AB 3770 hCoV-19/Italy/CAM-TIGEM-243/2020|EPI_ISL_776912|20 ···················· ·A·················· ···················· ···················· ···················· ····················
AB 3771 hCoV-19/Italy/CAM-TIGEM-235/2020|EPI_ISL_776913|20 ···················· ···················· ···················· ···················· ···················· ····················
AB 3772 hCoV-19/Italy/CAM-TIGEM-249/2020|EPI_ISL_776914|20 ···················· ···················· ···················· ···················· ···················· ····················
AB 3773 hCoV-19/Italy/CAM-TIGEM-198/2020|EPI_ISL_776915|20 ···················· ···················· ···················· ···················· ···················· ····················

| | | | | | | | |
|---|---|---|---|---|---|---|---|
| AB | 3774 hCoV-19/Italy/CAM-TIGEM-212/2020\|EPI_ISL_776916\|20 | ···················· | ···················· | ···················· | ···················· | ·····T·············· | ···················· |
| AB | 3775 hCoV-19/Italy/CAM-TIGEM-215/2020\|EPI_ISL_776917\|20 | ···················· | ···················· | ···················· | ···················· | ·····T·············· | ···················· |
| AB | 3776 hCoV-19/Italy/CAM-TIGEM-214/2020\|EPI_ISL_776918\|20 | ···················· | ·A·················· | ···················· | ···················· | ···················· | ···················· |
| AB | 3777 hCoV-19/Italy/CAM-TIGEM-209/2020\|EPI_ISL_776919\|20 | ···················· | ·A·················· | ···················· | ···················· | ···················· | ···················· |
| AB | 3778 hCoV-19/Italy/CAM-TIGEM-201/2020\|EPI_ISL_776920\|20 | ···················· | ···················· | ···················· | ···················· | ···················· | ···················· |
| AB | 3779 hCoV-19/Italy/CAM-TIGEM-290/2020\|EPI_ISL_776921\|20 | ···················· | ···················· | ···················· | ···················· | ···················· | ···················· |
| AB | 3780 hCoV-19/Italy/CAM-TIGEM-269/2020\|EPI_ISL_776922\|20 | ···················· | ···················· | ···················· | ···················· | ···················· | ···················· |
| AB | 3781 hCoV-19/Italy/CAM-TIGEM-254/2020\|EPI_ISL_776923\|20 | ···················· | ···················· | ···················· | ···················· | ···················· | ···················· |
| AB | 3782 hCoV-19/Italy/CAM-TIGEM-263/2020\|EPI_ISL_776924\|20 | ···················· | ···················· | ···················· | ···················· | ···················· | ···················· |
| AB | 3783 hCoV-19/Italy/CAM-TIGEM-273/2020\|EPI_ISL_776927\|20 | ···················· | ···················· | ···················· | ···················· | ···················· | ···················· |
| AB | 3784 hCoV-19/Italy/CAM-TIGEM-283/2020\|EPI_ISL_776929\|20 | ···················· | ···················· | ···················· | ···················· | ·····T·············· | ···················· |
| AB | 3785 hCoV-19/Italy/CAM-TIGEM-274/2020\|EPI_ISL_776930\|20 | ···················· | ···················· | ···················· | ···················· | ···················· | ···················· |
| AB | 3786 hCoV-19/Italy/CAM-TIGEM-265/2020\|EPI_ISL_776931\|20 | ···················· | ···················· | ···················· | ···················· | ···················· | ···················· |
| AB | 3787 hCoV-19/Italy/CAM-TIGEM-285/2020\|EPI_ISL_776943\|20 | ···················· | ···················· | ···················· | ···················· | ···················· | ···················· |
| AB | 3788 hCoV-19/Italy/CAM-TIGEM-268/2020\|EPI_ISL_776944\|20 | ···················· | ···················· | ···················· | ···················· | ···················· | ···················· |
| AB | 3789 hCoV-19/Italy/CAM-TIGEM-257/2020\|EPI_ISL_776945\|20 | ···················· | ···················· | ···················· | ···················· | ···················· | ···················· |
| AB | 3790 hCoV-19/Italy/CAM-TIGEM-264/2020\|EPI_ISL_776946\|20 | ···················· | ···················· | ···················· | ···················· | ···················· | ···················· |
| AB | 3791 hCoV-19/Italy/CAM-TIGEM-260/2020\|EPI_ISL_776947\|20 | ···················· | ···················· | ···················· | ···················· | ·····T·············· | ···················· |
| AB | 3792 hCoV-19/Italy/CAM-TIGEM-256/2020\|EPI_ISL_776948\|20 | ···················· | ···················· | ···················· | ···················· | ·····T·············· | ···················· |
| AB | 3793 hCoV-19/Italy/CAM-TIGEM-270/2020\|EPI_ISL_776949\|20 | ···················· | ···················· | ···················· | ···················· | ···················· | ···················· |
| AB | 3794 hCoV-19/Italy/CAM-TIGEM-291/2020\|EPI_ISL_776950\|20 | ···················· | ···················· | ···················· | ···················· | ·····T·············· | ···················· |
| AB | 3795 hCoV-19/Italy/CAM-TIGEM-296/2020\|EPI_ISL_776951\|20 | ···················· | ···················· | ···················· | ···················· | ···················· | ···················· |
| AB | 3796 hCoV-19/Italy/CAM-TIGEM-282/2020\|EPI_ISL_776952\|20 | ···················· | ···················· | ···················· | ···················· | ···················· | ···················· |
| AB | 3797 hCoV-19/Italy/CAM-TIGEM-276/2020\|EPI_ISL_776953\|20 | ···················· | ···················· | ···················· | ···················· | ···················· | ···················· |
| AB | 3798 hCoV-19/Italy/CAM-TIGEM-297/2020\|EPI_ISL_776954\|20 | ···················· | ···················· | ···················· | ···················· | ···················· | ···················· |
| AB | 3799 hCoV-19/Italy/CAM-TIGEM-277/2020\|EPI_ISL_776955\|20 | ···················· | ···················· | ···················· | ···················· | ···················· | ···················· |
| AB | 3800 hCoV-19/Italy/CAM-TIGEM-262/2020\|EPI_ISL_776956\|20 | ···················· | ···················· | ···················· | ···················· | ···················· | ···················· |
| AB | 3801 hCoV-19/Italy/CAM-TIGEM-275/2020\|EPI_ISL_776957\|20 | ···················· | ···················· | ···················· | ···················· | ···················· | ···················· |
| AB | 3802 hCoV-19/Italy/CAM-TIGEM-266/2020\|EPI_ISL_776958\|20 | ···················· | ·A·················· | ···················· | ···················· | ···················· | ···················· |
| AB | 3803 hCoV-19/Italy/CAM-TIGEM-267/2020\|EPI_ISL_776959\|20 | ···················· | ···················· | ···················· | ···················· | ···················· | ···················· |
| AB | 3804 hCoV-19/Italy/CAM-TIGEM-374/2020\|EPI_ISL_776960\|20 | ···················· | ···················· | ···················· | ···················· | ···················· | ···················· |
| AB | 3805 hCoV-19/Italy/CAM-TIGEM-368/2020\|EPI_ISL_776961\|20 | ···················· | ···················· | ···················· | ···················· | ···················· | ···················· |
| AB | 3806 hCoV-19/Italy/CAM-TIGEM-363/2020\|EPI_ISL_776963\|20 | ···················· | ···················· | ···················· | ···················· | ···················· | ···················· |
| AB | 3807 hCoV-19/Italy/CAM-TIGEM-312/2020\|EPI_ISL_776964\|20 | ···················· | ···················· | ···················· | ···················· | ·····T·············· | ···················· |
| AB | 3808 hCoV-19/Italy/CAM-TIGEM-359/2020\|EPI_ISL_776965\|20 | ···················· | ···················· | ···················· | ···················· | ·····T·············· | ···················· |
| AB | 3809 hCoV-19/Italy/CAM-TIGEM-304/2020\|EPI_ISL_776966\|20 | ···················· | ···················· | ···················· | ···················· | ·····T·············· | ···················· |
| AB | 3810 hCoV-19/Italy/CAM-TIGEM-325/2020\|EPI_ISL_776967\|20 | ···················· | ···················· | ···················· | ···················· | ·····T·············· | ···················· |
| AB | 3811 hCoV-19/Italy/CAM-TIGEM-305/2020\|EPI_ISL_776968\|20 | ···················· | ···················· | ···················· | ···················· | ···················· | ···················· |
| AB | 3812 hCoV-19/Italy/CAM-TIGEM-344/2020\|EPI_ISL_776969\|20 | ···················· | ···················· | ···················· | ···················· | ···················· | ···················· |
| AB | 3813 hCoV-19/Italy/CAM-TIGEM-324/2020\|EPI_ISL_776971\|20 | ···················· | ···················· | ···················· | ···················· | ···················· | ···················· |
| AB | 3814 hCoV-19/Italy/CAM-TIGEM-338/2020\|EPI_ISL_776973\|20 | ···················· | ···················· | ···················· | ···················· | ···················· | ···················· |
| AB | 3815 hCoV-19/Italy/CAM-TIGEM-341/2020\|EPI_ISL_776974\|20 | ···················· | ···················· | ···················· | ···················· | ···················· | ···················· |
| AB | 3816 hCoV-19/Italy/CAM-TIGEM-330/2020\|EPI_ISL_776975\|20 | ···················· | ···················· | ···················· | ···················· | ···················· | ···················· |
| AB | 3817 hCoV-19/Italy/CAM-TIGEM-350/2020\|EPI_ISL_776976\|20 | ···················· | ···················· | ···················· | ···················· | ···················· | ···················· |
| AB | 3818 hCoV-19/Italy/CAM-TIGEM-317/2020\|EPI_ISL_776977\|20 | ···················· | ···················· | ···················· | ···················· | ···················· | ···················· |
| AB | 3819 hCoV-19/Italy/CAM-TIGEM-300/2020\|EPI_ISL_776978\|20 | ···················· | ···················· | ···················· | ···················· | ···················· | ···················· |
| AB | 3820 hCoV-19/Italy/CAM-TIGEM-322/2020\|EPI_ISL_776980\|20 | ···················· | ···················· | ···················· | ···················· | ·····T·············· | ···················· |
| AB | 3821 hCoV-19/Italy/CAM-TIGEM-307/2020\|EPI_ISL_776981\|20 | ···················· | ···················· | ···················· | ···················· | ···················· | ···················· |
| AB | 3822 hCoV-19/Italy/CAM-TIGEM-345/2020\|EPI_ISL_776982\|20 | ···················· | ···················· | ···················· | ···················· | ·····T·············· | ···················· |
| AB | 3823 hCoV-19/Italy/CAM-TIGEM-352/2020\|EPI_ISL_776983\|20 | ···················· | ···················· | ···················· | ···················· | ···················· | ···················· |
| AB | 3824 hCoV-19/Italy/CAM-TIGEM-328/2020\|EPI_ISL_776984\|20 | ···················· | ···················· | ···················· | ···················· | ···················· | ···················· |
| AB | 3825 hCoV-19/Italy/CAM-TIGEM-334/2020\|EPI_ISL_776985\|20 | ···················· | ···················· | ···················· | ···················· | ···················· | ···················· |
| AB | 3826 hCoV-19/Italy/CAM-TIGEM-342/2020\|EPI_ISL_776986\|20 | ···················· | ···················· | ···················· | ···················· | ···················· | ···················· |
| AB | 3827 hCoV-19/Italy/CAM-TIGEM-348/2020\|EPI_ISL_776987\|20 | ···················· | ···················· | ···················· | ···················· | ···················· | ···················· |
| AB | 3828 hCoV-19/Italy/CAM-TIGEM-354/2020\|EPI_ISL_776988\|20 | ···················· | ···················· | ···················· | ···················· | ···················· | ···················· |
| AB | 3829 hCoV-19/Italy/CAM-TIGEM-313/2020\|EPI_ISL_776989\|20 | ···················· | ···················· | ···················· | ···················· | ···················· | ···················· |
| AB | 3830 hCoV-19/Italy/CAM-TIGEM-364/2020\|EPI_ISL_776998\|20 | ···················· | ···················· | ···················· | ···················· | ·····T·············· | ···················· |
| AB | 3831 hCoV-19/Italy/CAM-TIGEM-366/2020\|EPI_ISL_776999\|20 | ···················· | ···················· | ···················· | ···················· | ·····T·············· | ···················· |
| AB | 3832 hCoV-19/Brazil/RS-00641/2020\|EPI_ISL_779160\|2020-1 | ···················· | ···················· | ···················· | ···················· | ···················· | ···················· |
| AB | 3833 hCoV-19/Brazil/RS-00644/2020\|EPI_ISL_779161\|2020-1 | ···················· | ·····A·············· | ···················· | ···················· | ···················· | ···················· |
| AB | 3834 hCoV-19/Brazil/RS-00658/2020\|EPI_ISL_779162\|2020-1 | ···················· | ···················· | ···················· | ···················· | ···················· | ···················· |
| AB | 3835 hCoV-19/Brazil/RS-00664/2020\|EPI_ISL_779163\|2020-1 | ···················· | ·····A·············· | ···················· | ···················· | ···················· | ···················· |
| AB | 3836 hCoV-19/Brazil/RS-00671/2020\|EPI_ISL_779164\|2020-1 | ···················· | ···················· | ···················· | ···················· | ···················· | ···················· |
| AB | 3837 hCoV-19/Brazil/RS-00677/2020\|EPI_ISL_779165\|2020-1 | ···················· | ···················· | ···················· | ···················· | ···················· | ···················· |
| AB | 3838 hCoV-19/Brazil/RS-00678/2020\|EPI_ISL_779166\|2020-1 | ···················· | ···················· | ···················· | ···················· | ···················· | ···················· |
| AB | 3839 hCoV-19/Brazil/RS-00687/2020\|EPI_ISL_779167\|2020-1 | ···················· | ·····A·············· | ···················· | ···················· | ···················· | ···················· |
| AB | 3840 hCoV-19/Brazil/RS-00688/2020\|EPI_ISL_779168\|2020-1 | ···················· | ·····A·············· | ···················· | ···················· | ···················· | ···················· |
| AB | 3841 hCoV-19/England/ALDP-CB4263/2020\|EPI_ISL_761027\|20 | ···················· | ···················· | ···················· | ···················· | ··A················· | ···················· |
| AB | 3842 hCoV-19/England/QEUH-CADD4E/2020\|EPI_ISL_761028\|20 | ···················· | ···················· | ···················· | ···················· | ···················· | ···················· |
| AB | 3843 hCoV-19/England/ALDP-CB4E32/2020\|EPI_ISL_761029\|20 | ···················· | ···················· | ···················· | ···················· | ···················· | ···················· |
| AB | 3844 hCoV-19/England/ALDP-CB3DCD/2020\|EPI_ISL_761082\|20 | ···················· | ···················· | ···················· | ···················· | ·····T·············· | ···················· |
| AB | 3845 hCoV-19/England/ALDP-CB4D26/2020\|EPI_ISL_761084\|20 | ···················· | ···················· | ···················· | ···················· | ···················· | ···················· |
| AB | 3846 hCoV-19/England/ALDP-CB3E60/2020\|EPI_ISL_761085\|20 | ···················· | ···················· | ···················· | ···················· | ···················· | ···················· |
| AB | 3847 hCoV-19/England/ALDP-CB3871/2020\|EPI_ISL_761088\|20 | ···················· | ···················· | ···················· | ···················· | ···················· | ···················· |
| AB | 3848 hCoV-19/England/ALDP-CB45B5/2020\|EPI_ISL_761091\|20 | ···················· | ···················· | ···················· | ···················· | ·····T·············· | ···················· |
| AB | 3849 hCoV-19/England/ALDP-CB3914/2020\|EPI_ISL_761092\|20 | ···················· | ···················· | ···················· | ···················· | ···················· | ···················· |
| AB | 3850 hCoV-19/England/ALDP-CB67BD/2020\|EPI_ISL_761093\|20 | ···················· | ···················· | ···················· | ···················· | ···················· | ···················· |
| AB | 3851 hCoV-19/England/QEUH-CAD6E9/2020\|EPI_ISL_761094\|20 | ···················· | ···················· | ···················· | ···················· | ···················· | ···················· |
| AB | 3852 hCoV-19/England/QEUH-CADF39/2020\|EPI_ISL_761095\|20 | ···················· | ···················· | ···················· | ···················· | ···················· | ···················· |
| AB | 3853 hCoV-19/England/ALDP-CB3BC3/2020\|EPI_ISL_761096\|20 | ···················· | ···················· | ···················· | ···················· | ···················· | ···················· |

| | | | | | | | | |
|---|---|---|---|---|---|---|---|---|
| AB | 3854 | hCoV-19/England/ALDP-CB4D17/2020\|EPI_ISL_761097\|20 | ···················· | ···················· | ····················· | ···················· | ···················· | ···················· |
| AB | 3855 | hCoV-19/England/ALDP-CB5C64/2020\|EPI_ISL_761099\|20 | ···················· | ···················· | ····················· | ···················· | ···················· | ···················· |
| AB | 3856 | hCoV-19/Wales/ALDP-C39749/2020\|EPI_ISL_767200\|2020 | ···················· | ···················· | ····················· | ···················· | ···················· | ···················· |
| AB | 3857 | hCoV-19/Wales/ALDP-C3B45F/2020\|EPI_ISL_767201\|2020 | ···················· | ···················· | ····················· | ···················· | ···················· | ···················· |
| AB | 3858 | hCoV-19/Wales/ALDP-C3973A/2020\|EPI_ISL_767203\|2020 | ···················· | ···················· | ····················· | ···················· | ···················· | ···················· |
| AB | 3859 | hCoV-19/Wales/ALDP-C3B370/2020\|EPI_ISL_767204\|2020 | ···················· | ···················· | ····················· | ···················· | ···················· | ···················· |
| AB | 3860 | hCoV-19/Wales/ALDP-C3B440/2020\|EPI_ISL_767206\|2020 | ···················· | ···················· | ····················· | ···················· | ···················· | ···················· |
| AB | 3861 | hCoV-19/Wales/ALDP-C39D56/2020\|EPI_ISL_767207\|2020 | ···················· | ···················· | ····················· | ···················· | ···················· | ···················· |
| AB | 3862 | hCoV-19/Wales/ALDP-C3B3BC/2020\|EPI_ISL_767208\|2020 | ···················· | ···················· | ····················· | ···················· | ···················· | ···················· |
| AB | 3863 | hCoV-19/Wales/ALDP-C3B3CB/2020\|EPI_ISL_767209\|2020 | ···················· | ···················· | ····················· | ···················· | ···················· | ···················· |
| AB | 3864 | hCoV-19/Wales/ALDP-C3AE52/2020\|EPI_ISL_767210\|2020 | ···················· | ···················· | ····················· | ···················· | ···················· | ···················· |
| AB | 3865 | hCoV-19/Wales/ALDP-C3A4AB/2020\|EPI_ISL_767212\|2020 | ···················· | ···················· | ····················· | ···················· | ···················· | ···················· |
| AB | 3866 | hCoV-19/Wales/ALDP-C3A4F6/2020\|EPI_ISL_767213\|2020 | ···················· | ···················· | ····················· | ···················· | ···················· | ···················· |
| AB | 3867 | hCoV-19/Wales/ALDP-C398BF/2020\|EPI_ISL_767214\|2020 | ···················· | ···················· | ····················· | ···················· | ···················· | ···················· |
| AB | 3868 | hCoV-19/Wales/ALDP-C39758/2020\|EPI_ISL_767215\|2020 | ···················· | ···················· | ····················· | ···················· | ···················· | ···················· |
| AB | 3869 | hCoV-19/Wales/ALDP-C3BA6C/2020\|EPI_ISL_767216\|2020 | ···················· | ···················· | ····················· | ···················· | ···················· | ···················· |
| AB | 3870 | hCoV-19/Wales/ALDP-C3C4F4/2020\|EPI_ISL_767217\|2020 | ···················· | ······A············· | ····················· | ···················· | ···················· | ···················· |
| AB | 3871 | hCoV-19/Wales/ALDP-C3B905/2020\|EPI_ISL_767218\|2020 | ···················· | ···················· | ····················· | ···················· | ····T··············· | ···················· |
| AB | 3872 | hCoV-19/Wales/ALDP-C3CC29/2020\|EPI_ISL_767219\|2020 | ···················· | ···················· | ····················· | ···················· | ···················· | ···················· |
| AB | 3873 | hCoV-19/Wales/ALDP-C3CCED/2020\|EPI_ISL_767220\|2020 | ···················· | ···················· | ····················· | ···················· | ···················· | ···················· |
| AB | 3874 | hCoV-19/Wales/ALDP-C39E17/2020\|EPI_ISL_767221\|2020 | ···················· | ···················· | ····················· | ···················· | ···················· | ···················· |
| AB | 3875 | hCoV-19/Wales/ALDP-C3BA3F/2020\|EPI_ISL_767223\|2020 | ···················· | ···················· | ····················· | ···················· | ···················· | ···················· |
| AB | 3876 | hCoV-19/Wales/ALDP-C3C306/2020\|EPI_ISL_767224\|2020 | ···················· | ···················· | ····················· | ···················· | ···················· | ···················· |
| AB | 3877 | hCoV-19/Wales/ALDP-C3BB87/2020\|EPI_ISL_767225\|2020 | ···················· | ···················· | ····················· | ···················· | ···················· | ···A················ |
| AB | 3878 | hCoV-19/Wales/ALDP-C3BA99/2020\|EPI_ISL_767226\|2020 | ···················· | ···················· | ····················· | ···················· | ···················· | ···················· |
| AB | 3879 | hCoV-19/Wales/ALDP-C3B8F9/2020\|EPI_ISL_767229\|2020 | ···················· | ···················· | ····················· | ···················· | ···················· | ···················· |
| AB | 3880 | hCoV-19/Wales/ALDP-C3BBF0/2020\|EPI_ISL_767231\|2020 | ···················· | ······A············· | ····················· | ···················· | ···················· | ···················· |
| AB | 3881 | hCoV-19/Wales/ALDP-C3BA7B/2020\|EPI_ISL_767232\|2020 | ···················· | ···················· | ····················· | ···················· | ···················· | ···················· |
| AB | 3882 | hCoV-19/Wales/ALDP-C3BA8A/2020\|EPI_ISL_767233\|2020 | ···················· | ···················· | ····················· | ···················· | ···················· | ···················· |
| AB | 3883 | hCoV-19/Wales/ALDP-C3C2FA/2020\|EPI_ISL_767234\|2020 | ···················· | ···················· | ····················· | ···················· | ···················· | ···················· |
| AB | 3884 | hCoV-19/Wales/ALDP-C3C290/2020\|EPI_ISL_767235\|2020 | ···················· | ···················· | ····················· | ···················· | ···················· | ···················· |
| AB | 3885 | hCoV-19/Wales/ALDP-C3C2AF/2020\|EPI_ISL_767236\|2020 | ···················· | ···················· | ····················· | ···················· | ···················· | ···················· |
| AB | 3886 | hCoV-19/Wales/MILK-BB17E0/2020\|EPI_ISL_767239\|2020 | ···················· | ···················· | ····················· | ···················· | ···················· | ···················· |
| AB | 3887 | hCoV-19/Wales/ALDP-C2327E/2020\|EPI_ISL_767240\|2020 | ···················· | ···················· | ····················· | ···················· | ···················· | ···················· |
| AB | 3888 | hCoV-19/Wales/ALDP-C230DE/2020\|EPI_ISL_767241\|2020 | ···················· | ···················· | ····················· | ···················· | ···················· | ···················· |
| AB | 3889 | hCoV-19/Wales/ALDP-C22FE1/2020\|EPI_ISL_767242\|2020 | ···················· | ······A············· | ····················· | ···················· | ···················· | ···················· |
| AB | 3890 | hCoV-19/Wales/ALDP-C22FD2/2020\|EPI_ISL_767243\|2020 | ···················· | ···················· | ····················· | ···················· | ···················· | ···A················ |
| AB | 3891 | hCoV-19/Wales/ALDP-C22FA5/2020\|EPI_ISL_767244\|2020 | ···················· | ···················· | ····················· | ···················· | ···················· | ···················· |
| AB | 3892 | hCoV-19/Wales/ALDP-C22F78/2020\|EPI_ISL_767245\|2020 | ···················· | ···················· | ····················· | ···················· | ···················· | ···················· |
| AB | 3893 | hCoV-19/Wales/ALDP-C23180/2020\|EPI_ISL_767246\|2020 | ···················· | ···················· | ····················· | ···················· | ···················· | ···················· |
| AB | 3894 | hCoV-19/Wales/ALDP-C22F96/2020\|EPI_ISL_767247\|2020 | ···················· | ···················· | ····················· | ···················· | ···················· | ···················· |
| AB | 3895 | hCoV-19/Wales/ALDP-C22F87/2020\|EPI_ISL_767248\|2020 | ···················· | ······A············· | ····················· | ···················· | ···················· | ···················· |
| AB | 3896 | hCoV-19/Wales/ALDP-C231DB/2020\|EPI_ISL_767249\|2020 | ···················· | ···················· | ····················· | ···················· | ···················· | ···················· |
| AB | 3897 | hCoV-19/Wales/ALDP-C3A57B/2020\|EPI_ISL_767253\|2020 | ···················· | ···················· | ····················· | ···················· | ···················· | ···················· |
| AB | 3898 | hCoV-19/Wales/ALDP-C3A6C3/2020\|EPI_ISL_767261\|2020 | ···················· | ···················· | ····················· | ···················· | ···················· | ···················· |
| AB | 3899 | hCoV-19/Wales/ALDP-C3A669/2020\|EPI_ISL_767262\|2020 | ···················· | ···················· | ····················· | ···················· | ···················· | ···················· |
| AB | 3900 | hCoV-19/Wales/ALDP-C39819/2020\|EPI_ISL_767263\|2020 | ···················· | ···················· | ····················· | ···················· | ···················· | ···················· |
| AB | 3901 | hCoV-19/Wales/ALDP-C3911E/2020\|EPI_ISL_767264\|2020 | ···················· | ···················· | ····················· | ···················· | ···················· | ···················· |
| AB | 3902 | hCoV-19/Wales/ALDP-C39828/2020\|EPI_ISL_767265\|2020 | ···················· | ···················· | ····················· | ···················· | ···················· | ···················· |
| AB | 3903 | hCoV-19/Wales/ALDP-C3A53F/2020\|EPI_ISL_767268\|2020 | ···················· | ···················· | ····················· | ···················· | ···················· | ···················· |
| AB | 3904 | hCoV-19/Wales/ALDP-C39864/2020\|EPI_ISL_767269\|2020 | ···················· | ···················· | ····················· | ···················· | ···················· | ···················· |
| AB | 3905 | hCoV-19/Wales/ALDP-C395A9/2020\|EPI_ISL_767270\|2020 | ···················· | ···················· | ····················· | ···················· | ···················· | ···················· |
| AB | 3906 | hCoV-19/Wales/ALDP-C39846/2020\|EPI_ISL_767272\|2020 | ···················· | ···················· | ····················· | ···················· | ···················· | ···················· |
| AB | 3907 | hCoV-19/Wales/ALDP-C3A809/2020\|EPI_ISL_767273\|2020 | ···················· | ···················· | ····················· | ···················· | ···················· | ···················· |
| AB | 3908 | hCoV-19/Wales/ALDP-C3913C/2020\|EPI_ISL_767274\|2020 | ···················· | ···················· | ····················· | ···················· | ···················· | ···················· |
| AB | 3909 | hCoV-19/Wales/ALDP-C39178/2020\|EPI_ISL_767275\|2020 | ···················· | ···················· | ····················· | ···················· | ···················· | ···················· |
| AB | 3910 | hCoV-19/Wales/ALDP-C3958B/2020\|EPI_ISL_767276\|2020 | ···················· | ···················· | ····················· | ···················· | ···················· | ···················· |
| AB | 3911 | hCoV-19/Wales/ALDP-C3CFE4/2020\|EPI_ISL_767277\|2020 | ···················· | ······A············· | ····················· | ···················· | ···················· | ···················· |
| AB | 3912 | hCoV-19/Wales/ALDP-C3AC2B/2020\|EPI_ISL_767278\|2020 | ···················· | ···················· | ····················· | ···················· | ···················· | ···················· |
| AB | 3913 | hCoV-19/Wales/ALDP-C3BEF7/2020\|EPI_ISL_767279\|2020 | ···················· | ···················· | ····················· | ···················· | ···················· | ···················· |
| AB | 3914 | hCoV-19/Wales/ALDP-C3D280/2020\|EPI_ISL_767281\|2020 | ···················· | ···················· | ····················· | ···················· | ···················· | ···················· |
| AB | 3915 | hCoV-19/Wales/ALDP-C3BF03/2020\|EPI_ISL_767282\|2020 | ···················· | ···················· | ····················· | ···················· | ···················· | ···················· |
| AB | 3916 | hCoV-19/Wales/ALDP-C3AC3A/2020\|EPI_ISL_767284\|2020 | ···················· | ···················· | ····················· | ···················· | ···················· | ···················· |
| AB | 3917 | hCoV-19/Wales/ALDP-C3D39C/2020\|EPI_ISL_767285\|2020 | ···················· | ···················· | ····················· | ···················· | ···················· | ···················· |
| AB | 3918 | hCoV-19/Wales/ALDP-C3BEE8/2020\|EPI_ISL_767286\|2020 | ···················· | ···················· | ····················· | ···················· | ···················· | ···················· |
| AB | 3919 | hCoV-19/Wales/ALDP-C3BED9/2020\|EPI_ISL_767288\|2020 | ···················· | ···················· | ····················· | ···················· | ···················· | ···················· |
| AB | 3920 | hCoV-19/Wales/ALDP-C3D02C/2020\|EPI_ISL_767289\|2020 | ···················· | ···················· | ····················· | ···················· | ···················· | ···················· |
| AB | 3921 | hCoV-19/Wales/ALDP-C3D00E/2020\|EPI_ISL_767290\|2020 | ···················· | ···················· | ····················· | ···················· | ···················· | ···················· |
| AB | 3922 | hCoV-19/Wales/ALDP-C04B5A/2020\|EPI_ISL_767291\|2020 | ···················· | ···················· | ····················· | ···················· | ···················· | ···················· |
| AB | 3923 | hCoV-19/Wales/ALDP-C04E33/2020\|EPI_ISL_767292\|2020 | ···················· | ···················· | ····················· | ···················· | ···················· | ···················· |
| AB | 3924 | hCoV-19/Wales/ALDP-C04DAF/2020\|EPI_ISL_767293\|2020 | ···················· | ···················· | ····················· | ···················· | ···················· | ···················· |
| AB | 3925 | hCoV-19/Wales/ALDP-C04E7F/2020\|EPI_ISL_767294\|2020 | ···················· | ···················· | ····················· | ···················· | ···················· | ···················· |
| AB | 3926 | hCoV-19/Wales/ALDP-C04941/2020\|EPI_ISL_767295\|2020 | ···················· | ···················· | ····················· | ···················· | ···················· | ···················· |
| AB | 3927 | hCoV-19/Wales/ALDP-C04E8E/2020\|EPI_ISL_767296\|2020 | ···················· | ···················· | ····················· | ···················· | ···················· | ···················· |
| AB | 3928 | hCoV-19/Wales/ALDP-C04E06/2020\|EPI_ISL_767297\|2020 | ···················· | ···················· | ····················· | ···················· | ···················· | ···················· |
| AB | 3929 | hCoV-19/Wales/ALDP-C04DBE/2020\|EPI_ISL_767298\|2020 | ·A·················· | ···················· | ····················· | ···················· | ···················· | ···················· |
| AB | 3930 | hCoV-19/USA/LA-EVTL1231/2020\|EPI_ISL_768500\|2020-1 | ···················· | ·A·················· | ····················· | ···················· | ···················· | ···················· |
| AB | 3931 | hCoV-19/USA/LA-EVTL1232/2020\|EPI_ISL_768501\|2020-1 | ···················· | ···················· | ····················· | ···················· | ···················· | ···················· |
| AB | 3932 | hCoV-19/USA/LA-EVTL1233/2020\|EPI_ISL_768502\|2020-1 | ···················· | ···················· | ····················· | ···················· | ···················· | ···················· |
| AB | 3933 | hCoV-19/USA/LA-EVTL1234/2020\|EPI_ISL_768503\|2020-1 | ···················· | ···················· | ····················· | ···················· | ···················· | ···················· |

```
AB	3934 hCoV-19/USA/LA-EVTL1235/2020|EPI_ISL_768504|2020-1	····················	····················	····················	····················	····················	····················
AB	3935 hCoV-19/USA/LA-EVTL1236/2020|EPI_ISL_768505|2020-1	····················	····················	····················	····················	····················	····················
AB	3936 hCoV-19/USA/LA-EVTL1237/2020|EPI_ISL_768506|2020-1	····················	·A··················	····················	····················	····················	····················
AB	3937 hCoV-19/USA/LA-EVTL1238/2020|EPI_ISL_768507|2020-1	····················	····················	····················	····················	····················	····················
AB	3938 hCoV-19/USA/LA-EVTL1239/2020|EPI_ISL_768508|2020-1	····················	····················	····················	····················	····················	····················
AB	3939 hCoV-19/USA/LA-EVTL1240/2020|EPI_ISL_768509|2020-1	····················	····················	····················	····················	····················	····················
AB	3940 hCoV-19/USA/LA-EVTL1241/2020|EPI_ISL_768510|2020-1	····················	····················	····················	····················	····················	····················
AB	3941 hCoV-19/USA/LA-EVTL1242/2020|EPI_ISL_768511|2020-1	····················	····················	····················	····················	····················	····················
AB	3942 hCoV-19/USA/LA-EVTL1243/2020|EPI_ISL_768512|2020-1	····················	····················	····················	····················	····················	····················
AB	3943 hCoV-19/USA/LA-EVTL1244/2020|EPI_ISL_768513|2020-1	····················	·A··················	····················	····················	····················	····················
AB	3944 hCoV-19/USA/LA-EVTL1245/2020|EPI_ISL_768514|2020-1	····················	····················	····················	····················	····················	····················
AB	3945 hCoV-19/USA/LA-EVTL1246/2020|EPI_ISL_768515|2020-1	····················	···A················	····················	····················	····················	····················
AB	3946 hCoV-19/USA/LA-EVTL1247/2020|EPI_ISL_768516|2020-1	····················	····················	····················	····················	····················	····················
AB	3947 hCoV-19/USA/LA-EVTL1249/2020|EPI_ISL_768518|2020-1	····················	····················	····················	····················	····················	····················
AB	3948 hCoV-19/USA/LA-EVTL1250/2020|EPI_ISL_768519|2020-1	····················	····················	····················	····················	····················	····················
AB	3949 hCoV-19/USA/LA-EVTL1251/2020|EPI_ISL_768520|2020-1	····················	····················	····················	····················	····················	····················
AB	3950 hCoV-19/Thailand/Chonburi_SQ_KI0001/2020|EPI_ISL_7	····················	·A··················	····················	····················	····················	··A·················
AB	3951 hCoV-19/Thailand/Bangkok_29/2020|EPI_ISL_768522|20	····················	····················	····················	····················	····················	····················
AB	3952 hCoV-19/Thailand/Singburi_97/2020|EPI_ISL_768523|2	····················	·A··················	····················	····················	····················	··A·················
AB	3953 hCoV-19/USA/LA-EVTL1125/2020|EPI_ISL_768525|2020-1	····················	····················	····················	····················	····················	····················
AB	3954 hCoV-19/Thailand/Chiang Rai_SM/2020|EPI_ISL_768528	····················	·A··················	····················	····················	····················	··A·················
AB	3955 hCoV-19/Thailand/Chiang Rai_PY/2020|EPI_ISL_768529	····················	·A··················	····················	····················	····················	··A·················
AB	3956 hCoV-19/Thailand/Samut Sakorn_692/2020|EPI_ISL_768	····················	·A··················	····················	····················	····················	··A·················
AB	3957 hCoV-19/Thailand/Samut Sakorn_849/2020|EPI_ISL_768	····················	·A··················	····················	····················	····················	··A·················
AB	3958 hCoV-19/Thailand/Samut Sakorn_1235/2020|EPI_ISL_76	····················	·A··················	····················	····················	····················	··A·················
AB	3959 hCoV-19/Thailand/Samut Sakorn_26/2020|EPI_ISL_7685	····················	·A··················	····················	····················	····················	··A·················
AB	3960 hCoV-19/Thailand/Samut Sakorn_858/2020|EPI_ISL_768	····················	·A··················	····················	····················	····················	··A·················
AB	3961 hCoV-19/Thailand/Samut Sakorn_AS/2020|EPI_ISL_7685	····················	·A··················	····················	····················	····················	··A·················
AB	3962 hCoV-19/Thailand/Samut Sakorn_1184-19/2020|EPI_ISL	····················	·A··················	····················	····················	····················	··A·················
AB	3963 hCoV-19/Thailand/Samut Sakorn_SB232/2020|EPI_ISL_7	····················	·A··················	····················	····················	····················	··A·················
AB	3964 hCoV-19/Thailand/Samut Sakorn_RB0009/2020|EPI_ISL_	····················	·A··················	····················	····················	····················	··A·················
AB	3965 hCoV-19/Thailand/Samut Sakorn_657/2020|EPI_ISL_768	····················	·A··················	····················	····················	····················	··A·················
AB	3966 hCoV-19/Thailand/Samut Sakorn_894/2020|EPI_ISL_768	····················	·A··················	····················	····················	····················	··A·················
AB	3967 hCoV-19/Thailand/Samut Sakorn_887/2020|EPI_ISL_768	····················	·A··················	····················	····················	····················	··A·················
AB	3968 hCoV-19/Thailand/Samut Sakorn_27/2020|EPI_ISL_7685	····················	·A··················	····················	····················	····················	··A·················
AB	3969 hCoV-19/Thailand/Samut Sakorn_DB0013/2020|EPI_ISL_	····················	·A··················	····················	····················	····················	··A·················
AB	3970 hCoV-19/England/MILK-C5157E/2020|EPI_ISL_769800|20	····················	····················	····················	····················	····················	····················
AB	3971 hCoV-19/England/MILK-C53433/2020|EPI_ISL_769801|20	····················	····················	····················	····················	····················	····················
AB	3972 hCoV-19/England/QEUH-C4BBDF/2020|EPI_ISL_769802|20	····················	····················	····················	····················	····················	····················
AB	3973 hCoV-19/England/MILK-C53424/2020|EPI_ISL_769808|20	····················	····················	····················	····················	····················	····················
AB	3974 hCoV-19/England/MILK-C5167B/2020|EPI_ISL_769809|20	····················	····················	····················	····················	····················	····················
AB	3975 hCoV-19/England/MILK-C514AE/2020|EPI_ISL_769811|20	····················	····················	····················	····················	····················	····················
AB	3976 hCoV-19/England/QEUH-C4B53B/2020|EPI_ISL_769813|20	····················	····················	····················	····················	····················	····················
AB	3977 hCoV-19/England/MILK-C539CB/2020|EPI_ISL_769814|20	····················	····················	····················	····················	····················	····················
AB	3978 hCoV-19/England/MILK-C53846/2020|EPI_ISL_769816|20	····················	····················	····················	····················	····················	····················
AB	3979 hCoV-19/England/QEUH-C4B647/2020|EPI_ISL_769818|20	····················	····················	····················	····················	····················	····················
AB	3980 hCoV-19/England/MILK-C51972/2020|EPI_ISL_769819|20	····················	····················	····················	····················	·····T··············	····················
AB	3981 hCoV-19/England/MILK-C538DD/2020|EPI_ISL_769823|20	····················	····················	····················	····················	····················	····················
AB	3982 hCoV-19/England/MILK-C5398F/2020|EPI_ISL_769825|20	····················	····················	····················	····················	····················	····················
AB	3983 hCoV-19/England/MILK-C539DA/2020|EPI_ISL_769827|20	····················	····················	····················	····················	····················	····················
AB	3984 hCoV-19/England/MILK-C5168A/2020|EPI_ISL_769828|20	····················	····················	····················	····················	····················	····················
AB	3985 hCoV-19/England/QEUH-C4B823/2020|EPI_ISL_769836|20	····················	····················	····················	····················	····················	····················
AB	3986 hCoV-19/England/QEUH-C4B8C9/2020|EPI_ISL_769837|20	····················	····················	····················	····················	····················	····················
AB	3987 hCoV-19/England/QEUH-C4B7CC/2020|EPI_ISL_769842|20	····················	····················	····················	····················	····················	····················
AB	3988 hCoV-19/England/MILK-C4FE0F/2020|EPI_ISL_769843|20	····················	·A··················	····················	····················	····················	··A·················
AB	3989 hCoV-19/England/MILK-C516A8/2020|EPI_ISL_769844|20	····················	····················	····················	····················	····················	····················
AB	3990 hCoV-19/England/MILK-C4FF2A/2020|EPI_ISL_769849|20	····················	····················	····················	····················	····················	····················
AB	3991 hCoV-19/England/QEUH-C4B9E4/2020|EPI_ISL_769850|20	····················	····················	····················	····················	·····T··············	····················
AB	3992 hCoV-19/England/MILK-C4FDE4/2020|EPI_ISL_769854|20	····················	····················	····················	····················	····················	····················
AB	3993 hCoV-19/England/MILK-C51954/2020|EPI_ISL_769855|20	····················	····················	····················	····················	····················	····················
AB	3994 hCoV-19/England/MILK-C53767/2020|EPI_ISL_769860|20	····················	····················	····················	····················	····················	····················
AB	3995 hCoV-19/England/QEUH-C4BB93/2020|EPI_ISL_769861|20	····················	····················	····················	····················	····················	····················
AB	3996 hCoV-19/England/QEUH-C4B7DB/2020|EPI_ISL_769862|20	····················	····················	····················	····················	····················	····················
AB	3997 hCoV-19/Morocco/FMP-42/2020|EPI_ISL_769863|2020-12	····················	····················	····················	····················	····················	····················
AB	3998 hCoV-19/England/205041300/2020|EPI_ISL_769868|2020	····················	····················	····················	····················	····················	····················
AB	3999 hCoV-19/England/205081145/2020|EPI_ISL_769869|2020	····················	····················	····················	····················	····················	····················
AB	4000 hCoV-19/Gibraltar/205121254/2020|EPI_ISL_769872|20	····················	·A··················	····················	····················	····················	····················
AB	4001 hCoV-19/England/205241494/2020|EPI_ISL_769876|2020	····················	····················	····················	····················	····················	····················
AB	4002 hCoV-19/England/205090690/2020|EPI_ISL_769878|2020	····················	····················	····················	····················	····················	····················
AB	4003 hCoV-19/USA/WY-WPHL-20063179/2020|EPI_ISL_769879|2	····················	····················	····················	····················	····················	····················
AB	4004 hCoV-19/USA/WY-WPHL-20063117/2020|EPI_ISL_769880|2	····················	····················	····················	····················	····················	····················
AB	4005 hCoV-19/USA/WY-WPHL-20063101/2020|EPI_ISL_769881|2	····················	····················	····················	····················	····················	····················
AB	4006 hCoV-19/USA/WY-WPHL-20063043/2020|EPI_ISL_769882|2	····················	··C·················	····················	····················	····················	····················
AB	4007 hCoV-19/USA/WY-WPHL-20063014/2020|EPI_ISL_769883|2	····················	····················	····················	····················	····················	····················
AB	4008 hCoV-19/USA/WY-WPHL-20062990/2020|EPI_ISL_769884|2	····················	····················	····················	····················	····················	····················
AB	4009 hCoV-19/USA/WY-WPHL-20062989/2020|EPI_ISL_769885|2	····················	····················	····················	····················	····················	····················
AB	4010 hCoV-19/USA/WY-WPHL-20062882/2020|EPI_ISL_769886|2	····················	·A··················	····················	····················	····················	····················
AB	4011 hCoV-19/USA/WY-WPHL-20062881/2020|EPI_ISL_769887|2	····················	··C·················	····················	····················	····················	····················
AB	4012 hCoV-19/USA/WY-WPHL-20062878/2020|EPI_ISL_769888|2	····················	·A··················	····················	····················	····················	····················
AB	4013 hCoV-19/USA/WY-WPHL-20062877/2020|EPI_ISL_769889|2	····················	··C·················	····················	····················	····················	····················
```

| | | | | | | | |
|---|---|---|---|---|---|---|---|
| AB | 4014 hCoV-19/USA/WY-WPHL-20062765/2020\|EPI_ISL_769890\|2 | ···················· | ···················· | ···················· | ···················· | ···················· | ···················· |
| AB | 4015 hCoV-19/USA/WY-WPHL-20062716/2020\|EPI_ISL_769891\|2 | ···················· | ···················· | ···················· | ···················· | ···················· | ···················· |
| AB | 4016 hCoV-19/USA/WY-WPHL-20062588/2020\|EPI_ISL_769892\|2 | ···················· | ·A·················· | ···················· | ···················· | ···················· | ···················· |
| AB | 4017 hCoV-19/USA/WY-WPHL-20062464/2020\|EPI_ISL_769893\|2 | ···················· | ···················· | ···················· | ···················· | ···················· | ···················· |
| AB | 4018 hCoV-19/USA/WY-WPHL-20062312/2020\|EPI_ISL_769894\|2 | ···················· | ···················· | ···················· | ···················· | ···················· | ···················· |
| AB | 4019 hCoV-19/USA/WY-WPHL-20062182/2020\|EPI_ISL_769895\|2 | ···················· | ···················· | ···················· | ···················· | ···················· | ···················· |
| AB | 4020 hCoV-19/USA/WY-WPHL-20062180/2020\|EPI_ISL_769896\|2 | ···················· | ···················· | ···················· | ···················· | ···················· | ···················· |
| AB | 4021 hCoV-19/USA/WY-WPHL-20062171/2020\|EPI_ISL_769897\|2 | ···················· | ·A·················· | ···················· | ···················· | ···················· | ···················· |
| AB | 4022 hCoV-19/USA/WY-WPHL-20062139/2020\|EPI_ISL_769898\|2 | ···················· | ···················· | ···················· | ···················· | ···················· | ···················· |
| AB | 4023 hCoV-19/USA/WY-WPHL-20061980/2020\|EPI_ISL_769899\|2 | ···················· | ···················· | ···················· | ···················· | ···················· | ···················· |
| AB | 4024 hCoV-19/USA/MN-MDH-2226/2020\|EPI_ISL_770800\|2020-1 | ···················· | ···················· | ···················· | ···················· | ···················· | ···················· |
| AB | 4025 hCoV-19/USA/MN-MDH-2227/2020\|EPI_ISL_770801\|2020-1 | ···················· | ···················· | ···················· | ···················· | ···················· | ···················· |
| AB | 4026 hCoV-19/USA/MN-MDH-2228/2020\|EPI_ISL_770802\|2020-1 | ···················· | ···················· | ···················· | ···················· | ···················· | ···················· |
| AB | 4028 hCoV-19/USA/MN-MDH-2230/2020\|EPI_ISL_770804\|2020-1 | ···················· | ···················· | ···················· | ···················· | ···················· | ···················· |
| AB | 4029 hCoV-19/USA/MN-MDH-2232/2020\|EPI_ISL_770806\|2020-1 | ···················· | ···················· | ···················· | ···················· | ···················· | ···················· |
| AB | 4030 hCoV-19/USA/MN-MDH-2233/2020\|EPI_ISL_770807\|2020-1 | ···················· | ···················· | ···················· | ···················· | ···················· | ···················· |
| AB | 4031 hCoV-19/USA/MN-MDH-2234/2020\|EPI_ISL_770808\|2020-1 | ···················· | ·A·················· | ···················· | ···················· | ···················· | ···················· |
| AB | 4032 hCoV-19/USA/MN-MDH-2235/2020\|EPI_ISL_770809\|2020-1 | ···················· | ·A·················· | ···················· | ···················· | ···················· | ···················· |
| AB | 4033 hCoV-19/USA/MN-MDH-2236/2020\|EPI_ISL_770810\|2020-1 | ···················· | ·A·················· | ···················· | ···················· | ···················· | ···················· |
| AB | 4034 hCoV-19/USA/MN-MDH-2237/2020\|EPI_ISL_770811\|2020-1 | ···················· | ···················· | ···················· | ···················· | ···················· | ···················· |
| AB | 4035 hCoV-19/USA/MN-MDH-2238/2020\|EPI_ISL_770812\|2020-1 | ···················· | ·A·················· | ···················· | ···················· | ···················· | ···················· |
| AB | 4036 hCoV-19/USA/MN-MDH-2239/2020\|EPI_ISL_770813\|2020-1 | ···················· | ···················· | ···················· | ···················· | ···················· | ···················· |
| AB | 4037 hCoV-19/USA/MN-MDH-2240/2020\|EPI_ISL_770814\|2020-1 | ···················· | ···················· | ···················· | ···················· | ···················· | ···················· |
| AB | 4038 hCoV-19/USA/MN-MDH-2241/2020\|EPI_ISL_770815\|2020-1 | ···················· | ···················· | ···················· | ···················· | ···················· | ···················· |
| AB | 4039 hCoV-19/USA/MN-MDH-2242/2020\|EPI_ISL_770816\|2020-1 | ···················· | ···················· | ···················· | ···················· | ···················· | ···················· |
| AB | 4040 hCoV-19/USA/MN-MDH-2243/2020\|EPI_ISL_770817\|2020-1 | ···················· | ···················· | ···················· | ···················· | ···················· | ···················· |
| AB | 4041 hCoV-19/USA/MN-MDH-2244/2020\|EPI_ISL_770818\|2020-1 | ···················· | ···················· | ···················· | ···················· | ···················· | ···················· |
| AB | 4042 hCoV-19/USA/MN-MDH-2245/2020\|EPI_ISL_770819\|2020-1 | ···················· | ···················· | ···················· | ···················· | ···················· | ···················· |
| AB | 4043 hCoV-19/Australia/NSW1378/2020\|EPI_ISL_770820\|2020 | ···················· | ···················· | ···················· | ···················· | ···················· | ···················· |
| AB | 4044 hCoV-19/Canada/ON-E70b/2020\|EPI_ISL_770823\|2020-03 | ···················· | ···················· | ···················· | ···················· | ···················· | ···················· |
| AB | 4045 hCoV-19/Canada/ON-E72b/2020\|EPI_ISL_770825\|2020-04 | ···················· | ···················· | ···················· | ···················· | ···················· | ···················· |
| AB | 4046 hCoV-19/Canada/ON-E74b/2020\|EPI_ISL_770826\|2020-10 | ···················· | ···················· | ···················· | ···················· | ···················· | ···················· |
| AB | 4047 hCoV-19/Canada/ON-E76b/2020\|EPI_ISL_770827\|2020-05 | ···················· | ···················· | ···················· | ···················· | ···················· | ···················· |
| AB | 4048 hCoV-19/Canada/ON-S1453/2020\|EPI_ISL_770830\|2020-0 | ···················· | ·A·················· | ···················· | ···················· | ···················· | ··A················· |
| AB | 4049 hCoV-19/Canada/ON-S1459/2020\|EPI_ISL_770832\|2020-0 | ···················· | ···················· | ···················· | ···················· | ···················· | ···················· |
| AB | 4050 hCoV-19/Canada/ON-S1460/2020\|EPI_ISL_770833\|2020-0 | ···················· | ···················· | ···················· | ···················· | ···················· | ···················· |
| AB | 4051 hCoV-19/Canada/ON-S1461/2020\|EPI_ISL_770834\|2020-0 | ···················· | ···················· | ···················· | ···················· | ···················· | ···················· |
| AB | 4052 hCoV-19/Canada/ON-S1462/2020\|EPI_ISL_770835\|2020-0 | ···················· | ···················· | ···················· | ···················· | ···················· | ···················· |
| AB | 4053 hCoV-19/Canada/ON-S1463/2020\|EPI_ISL_770836\|2020-0 | ···················· | ···················· | ···················· | ···················· | ···················· | ···················· |
| AB | 4054 hCoV-19/Canada/ON-S1464/2020\|EPI_ISL_770837\|2020-0 | ···················· | ···················· | ···················· | ···················· | ···················· | ···················· |
| AB | 4055 hCoV-19/Canada/ON-S1467/2020\|EPI_ISL_770838\|2020-0 | ···················· | ·A·················· | ···················· | ···················· | ···················· | ··A················· |
| AB | 4056 hCoV-19/Canada/ON-S1486/2020\|EPI_ISL_770839\|2020-0 | ···················· | ···················· | ···················· | ···················· | ···················· | ···················· |
| AB | 4057 hCoV-19/Canada/ON-S1487/2020\|EPI_ISL_770840\|2020-0 | ···················· | ···················· | ···················· | ···················· | ···················· | ···················· |
| AB | 4058 hCoV-19/Canada/ON-S1489/2020\|EPI_ISL_770841\|2020-0 | ···················· | ···················· | ···················· | ···················· | ···················· | ···················· |
| AB | 4059 hCoV-19/Canada/ON-S1490/2020\|EPI_ISL_770842\|2020-0 | ···················· | ···················· | ···················· | ···················· | ···················· | ···················· |
| AB | 4060 hCoV-19/Canada/ON-S1491/2020\|EPI_ISL_770843\|2020-0 | ···················· | ···················· | ···················· | ···················· | ···················· | ···················· |
| AB | 4061 hCoV-19/Canada/ON-S1493/2020\|EPI_ISL_770844\|2020-0 | ···················· | ·A·················· | ···················· | ···················· | ···················· | ··A················· |
| AB | 4062 hCoV-19/Luxembourg/LNS7364988/2020\|EPI_ISL_770878\| | ···················· | ···················· | ···················· | ···················· | ···················· | ···················· |
| AB | 4063 hCoV-19/Luxembourg/LNS3837315/2020\|EPI_ISL_770879\| | ···················· | ···················· | ···················· | ···················· | ···················· | ···················· |
| AB | 4064 hCoV-19/Luxembourg/LNS5049262/2020\|EPI_ISL_770893\| | ···················· | ···················· | ···················· | ···················· | ···················· | ···················· |
| AB | 4065 hCoV-19/Luxembourg/LNS0287451/2020\|EPI_ISL_770894\| | ···················· | ···················· | ···················· | ···················· | ···················· | ···················· |
| AB | 4066 hCoV-19/Luxembourg/LNS9744887/2020\|EPI_ISL_770895\| | ···················· | ···················· | ···················· | ···················· | ···················· | ···················· |
| AB | 4067 hCoV-19/Luxembourg/LNS0257136/2020\|EPI_ISL_770896\| | ···················· | ···················· | ···················· | ···················· | ···················· | ···················· |
| AB | 4068 hCoV-19/Luxembourg/LNS0224787/2020\|EPI_ISL_770897\| | ···················· | ···················· | ···················· | ···················· | ···················· | ···················· |
| AB | 4069 hCoV-19/Luxembourg/LNS6172039/2020\|EPI_ISL_770898\| | ···················· | ···················· | ···················· | ···················· | ···················· | ···················· |
| AB | 4070 hCoV-19/Luxembourg/LNS3817100/2020\|EPI_ISL_770899\| | ···················· | ···················· | ···················· | ···················· | ···················· | ···················· |
| AB | 4071 hCoV-19/Italy/CAM-TIGEM-306/2020\|EPI_ISL_776990\|20 | ···················· | ···················· | ···················· | ···················· | ···················· | ···················· |
| AB | 4072 hCoV-19/Italy/CAM-TIGEM-346/2020\|EPI_ISL_776991\|20 | ···················· | ···················· | ···················· | ···················· | ······T············· | ···················· |
| AB | 4073 hCoV-19/Italy/CAM-TIGEM-361/2020\|EPI_ISL_776992\|20 | ···················· | ···················· | ···················· | ···················· | ······T············· | ···················· |
| AB | 4074 hCoV-19/Italy/CAM-TIGEM-360/2020\|EPI_ISL_776993\|20 | ···················· | ···················· | ···················· | ···················· | ···················· | ···················· |
| AB | 4075 hCoV-19/USA/OH-ODH-SC35430/2020\|EPI_ISL_776994\|202 | ···················· | ···················· | ···················· | ···················· | ···················· | ···················· |
| AB | 4076 hCoV-19/Italy/CAM-TIGEM-379/2020\|EPI_ISL_776995\|20 | ···················· | ···················· | ···················· | ···················· | ······T············· | ···················· |
| AB | 4077 hCoV-19/Italy/CAM-TIGEM-332/2020\|EPI_ISL_776996\|20 | ···················· | ···················· | ···················· | ···················· | ······T············· | ···················· |
| AB | 4078 hCoV-19/Italy/CAM-TIGEM-365/2020\|EPI_ISL_776997\|20 | ···················· | ···················· | ···················· | ···················· | ······T············· | ···················· |
| AB | 4079 hCoV-19/Pakistan/JRCGR-KHI05/2020\|EPI_ISL_779255\|2 | ···················· | ···················· | ···················· | ···················· | ···················· | ···················· |
| AB | 4080 hCoV-19/Pakistan/JRCGR-KHI06/2020\|EPI_ISL_779256\|2 | ···················· | ···················· | ···················· | ···················· | ···················· | ···················· |
| AB | 4081 hCoV-19/Pakistan/JRCGR-KHI22/2020\|EPI_ISL_779257\|2 | ···················· | ···················· | ···················· | ···················· | ···················· | ···················· |
| AB | 4082 hCoV-19/Pakistan/JRCGR-KHI07/2020\|EPI_ISL_779258\|2 | ···················· | ···················· | ···················· | ···················· | ···················· | ···················· |
| AB | 4083 hCoV-19/Pakistan/JRCGR-KHI26/2020\|EPI_ISL_779260\|2 | ···················· | ·A·················· | ···················· | ···················· | ···················· | ··A················· |
| AB | 4084 hCoV-19/Pakistan/JRCGR-KHI28/2020\|EPI_ISL_779262\|2 | ···················· | ·A·················· | ···················· | ···················· | ···················· | ··A················· |
| AB | 4085 hCoV-19/Pakistan/JRCGR-KHI04/2020\|EPI_ISL_779264\|2 | ···················· | ···················· | ···················· | ···················· | ···················· | ···················· |
| AB | 4086 hCoV-19/Pakistan/JRCGR-KHI29/2020\|EPI_ISL_779265\|2 | ···················· | ·A·················· | ···················· | ···················· | ···················· | ··A················· |
| AB | 4087 hCoV-19/Pakistan/JRCGR-KHI30/2020\|EPI_ISL_779266\|2 | ···················· | ···················· | ···················· | ···················· | ···················· | ···················· |
| AB | 4088 hCoV-19/Pakistan/JRCGR-KHI40/2020\|EPI_ISL_779267\|2 | ···················· | ···················· | ···················· | ···················· | ···················· | ···················· |
| AB | 4089 hCoV-19/Pakistan/JRCGR-KHI03/2020\|EPI_ISL_779268\|2 | ···················· | ···················· | ···················· | ···················· | ···················· | ···················· |
| AB | 4090 hCoV-19/Pakistan/JRCGR-KHI02/2020\|EPI_ISL_779269\|2 | ···················· | ·A·················· | ···················· | ···················· | ···················· | ··A················· |
| AB | 4091 hCoV-19/Pakistan/JRCGR-KHI12/2020\|EPI_ISL_779271\|2 | ···················· | ···················· | ···················· | ···················· | ···················· | ···················· |
| AB | 4092 hCoV-19/Pakistan/JRCGR-KHI18/2020\|EPI_ISL_779272\|2 | ···················· | ···················· | ···················· | ···················· | ···················· | ···················· |
| AB | 4093 hCoV-19/Pakistan/JRCGR-KHI15/2020\|EPI_ISL_779275\|2 | ···················· | ···················· | ···················· | ···················· | ···················· | ···················· |
| AB | 4094 hCoV-19/Pakistan/JRCGR-KHI20/2020\|EPI_ISL_779276\|2 | ···················· | ···················· | ···················· | ···················· | ···················· | ···················· |

| | | | | | | | |
|---|---|---|---|---|---|---|---|
| AB | 4095 hCoV-19/Pakistan/JRCGR-KHI35/2020\|EPI_ISL_779277\|2 | ···················· | ·A·················· | ···················· | ···················· | ···················· | ···A················ |
| AB | 4096 hCoV-19/Pakistan/JRCGR-KHI11/2020\|EPI_ISL_779279\|2 | ···················· | ···················· | ···················· | ···················· | ···················· | ···················· |
| AB | 4097 hCoV-19/England/ALDP-CB6911/2020\|EPI_ISL_761100\|20 | ···················· | ·A·················· | ···················· | ···················· | ···················· | ···A················ |
| AB | 4098 hCoV-19/England/ALDP-CB4CCF/2020\|EPI_ISL_761101\|20 | ···················· | ···················· | ···················· | ···················· | ···················· | ···················· |
| AB | 4099 hCoV-19/England/ALDP-CB3E8E/2020\|EPI_ISL_761102\|20 | ···················· | ···················· | ···················· | ···················· | ···················· | ···················· |
| AB | 4100 hCoV-19/England/ALDP-CB3DDC/2020\|EPI_ISL_761103\|20 | ···················· | ···················· | ···················· | ···················· | ···················· | ···················· |
| AB | 4101 hCoV-19/England/ALDP-CB3826/2020\|EPI_ISL_761104\|20 | ···················· | ···················· | ···················· | ···················· | ···················· | ···················· |
| AB | 4102 hCoV-19/England/ALDP-CB4CB0/2020\|EPI_ISL_761106\|20 | ···················· | ···················· | ···················· | ···················· | ···················· | ···················· |
| AB | 4103 hCoV-19/England/ALDP-CB35B6/2020\|EPI_ISL_761108\|20 | ···················· | ···················· | ···················· | ···················· | ···················· | ···················· |
| AB | 4104 hCoV-19/England/ALDP-CB3510/2020\|EPI_ISL_761109\|20 | ···················· | ···················· | ···················· | ···················· | ···················· | ···················· |
| AB | 4105 hCoV-19/England/ALDP-CB403C/2020\|EPI_ISL_761111\|20 | ···················· | ···················· | ···················· | ···················· | ··A················· | ···················· |
| AB | 4106 hCoV-19/England/QEUH-CADB9F/2020\|EPI_ISL_761113\|20 | ···················· | ···················· | ···················· | ···················· | ···················· | ···················· |
| AB | 4107 hCoV-19/England/MILK-CA9CDC/2020\|EPI_ISL_762402\|20 | ···················· | ···················· | ···················· | ···················· | ···················· | ···················· |
| AB | 4108 hCoV-19/England/MILK-CA9AA5/2020\|EPI_ISL_762403\|20 | ···················· | ···················· | ···················· | ···················· | ···················· | ···················· |
| AB | 4109 hCoV-19/England/MILK-CA5159/2020\|EPI_ISL_762407\|20 | ···················· | ···················· | ···················· | ···················· | ·····T·············· | ···················· |
| AB | 4110 hCoV-19/England/MILK-CA94B6/2020\|EPI_ISL_762412\|20 | ···················· | ···················· | ···················· | ···················· | ·····T·············· | ···················· |
| AB | 4111 hCoV-19/England/MILK-CA4BC5/2020\|EPI_ISL_762416\|20 | ···················· | ···················· | ···················· | ···················· | ···················· | ···················· |
| AB | 4112 hCoV-19/England/MILK-CA455E/2020\|EPI_ISL_762418\|20 | ···················· | ···················· | ···················· | ···················· | ···················· | ···················· |
| AB | 4113 hCoV-19/England/MILK-CA49F8/2020\|EPI_ISL_762423\|20 | ···················· | ·A·················· | ···················· | ···················· | ···················· | ···A················ |
| AB | 4114 hCoV-19/England/MILK-CA4B5C/2020\|EPI_ISL_762436\|20 | ···················· | ···················· | ···················· | ···················· | ···················· | ···················· |
| AB | 4115 hCoV-19/England/MILK-CA4C86/2020\|EPI_ISL_762439\|20 | ···················· | ···················· | ···················· | ···················· | ···················· | ···················· |
| AB | 4116 hCoV-19/England/MILK-CA420C/2020\|EPI_ISL_762443\|20 | ···················· | ···················· | ···················· | ···················· | ···················· | ···················· |
| AB | 4117 hCoV-19/England/MILK-CA9D42/2020\|EPI_ISL_762444\|20 | ···················· | ···················· | ···················· | ···················· | ···················· | ···················· |
| AB | 4118 hCoV-19/England/CAMB-1BD5E4/2020\|EPI_ISL_764272\|20 | ···················· | ···················· | ···················· | ···················· | ···················· | ···················· |
| AB | 4122 hCoV-19/Wales/PHWC-4A5FB1/2020\|EPI_ISL_764282\|2020 | ···················· | ···················· | ···················· | ···················· | ···················· | ···················· |
| AB | 4123 hCoV-19/Wales/PHWC-4A61AB/2020\|EPI_ISL_764283\|2020 | ···················· | ···················· | ···················· | ···················· | ···················· | ···················· |
| AB | 4124 hCoV-19/England/CAMB-1BC62E/2020\|EPI_ISL_764284\|20 | ···················· | ···················· | ···················· | ···················· | ···················· | ···················· |
| AB | 4125 hCoV-19/England/CAMB-1BC6D3/2020\|EPI_ISL_764285\|20 | ···················· | ···················· | ···················· | ···················· | ···················· | ···················· |
| AB | 4126 hCoV-19/England/CAMB-1BC891/2020\|EPI_ISL_764286\|20 | ···················· | ···················· | ···················· | ···················· | ···················· | ···················· |
| AB | 4127 hCoV-19/England/CAMB-1BC8A0/2020\|EPI_ISL_764287\|20 | ···················· | ···················· | ···················· | ···················· | ···················· | ···················· |
| AB | 4128 hCoV-19/England/CAMB-1BC8BF/2020\|EPI_ISL_764288\|20 | ···················· | ···················· | ···················· | ···················· | ···················· | ···················· |
| AB | 4129 hCoV-19/England/CAMB-1BC8EC/2020\|EPI_ISL_764289\|20 | ···················· | ···················· | ···················· | ···················· | ·····T·············· | ···················· |
| AB | 4130 hCoV-19/England/CAMB-1BCA8C/2020\|EPI_ISL_764290\|20 | ···················· | ···················· | ···················· | ···················· | ···················· | ···················· |
| AB | 4131 hCoV-19/England/CAMB-1BCAD7/2020\|EPI_ISL_764292\|20 | ···················· | ···················· | ···················· | ···················· | ·····T·············· | ···················· |
| AB | 4132 hCoV-19/England/CAMB-1BCB01/2020\|EPI_ISL_764293\|20 | ···················· | ···················· | ···················· | ···················· | ···················· | ···················· |
| AB | 4133 hCoV-19/England/CAMB-1BCB10/2020\|EPI_ISL_764294\|20 | ···················· | ···················· | ···················· | ···················· | ···················· | ···················· |
| AB | 4134 hCoV-19/England/CAMB-1BCB4D/2020\|EPI_ISL_764295\|20 | ···················· | ···················· | ···················· | ···················· | ···················· | ···················· |
| AB | 4135 hCoV-19/USA/NY-Wadsworth-213737-01/2020\|EPI_ISL_76 | ···················· | ···················· | ···················· | ···················· | ···················· | ···················· |
| AB | 4136 hCoV-19/USA/NY-Wadsworth-213738-01/2020\|EPI_ISL_76 | ···················· | ···················· | ···················· | ···················· | ···················· | ···················· |
| AB | 4137 hCoV-19/USA/NY-Wadsworth-214713-01/2020\|EPI_ISL_76 | ···················· | ·A·················· | ···················· | ···················· | ···················· | ···················· |
| AB | 4138 hCoV-19/USA/NY-Wadsworth-215771-01/2020\|EPI_ISL_76 | ···················· | ···················· | ···················· | ···················· | ···················· | ···················· |
| AB | 4139 hCoV-19/USA/NY-Wadsworth-216883-01/2020\|EPI_ISL_76 | ···················· | ·A·················· | ···················· | ···················· | ···················· | ···················· |
| AB | 4140 hCoV-19/USA/NY-Wadsworth-218073-01/2020\|EPI_ISL_76 | ···················· | ·A·················· | ···················· | ···················· | ···················· | ···················· |
| AB | 4141 hCoV-19/USA/NY-Wadsworth-217616-01/2020\|EPI_ISL_76 | ···················· | ·A·················· | ···················· | ···················· | ···················· | ···················· |
| AB | 4142 hCoV-19/USA/NY-Wadsworth-217623-01/2020\|EPI_ISL_76 | ···················· | ···················· | ···················· | ···················· | ···················· | ···················· |
| AB | 4143 hCoV-19/USA/NY-Wadsworth-220236-01/2020\|EPI_ISL_76 | ···················· | ·A·················· | ···················· | ···················· | ···················· | ···················· |
| AB | 4144 hCoV-19/USA/NY-Wadsworth-222417-01/2020\|EPI_ISL_76 | ···················· | ···················· | ···················· | ···················· | ···················· | ···················· |
| AB | 4145 hCoV-19/USA/NY-Wadsworth-222415-01/2020\|EPI_ISL_76 | ···················· | ···················· | ···················· | ···················· | ···················· | ···················· |
| AB | 4146 hCoV-19/USA/NY-Wadsworth-223805-01/2020\|EPI_ISL_76 | ···················· | ···················· | ···················· | ···················· | ···················· | ···················· |
| AB | 4147 hCoV-19/USA/NY-Wadsworth-223775-01/2020\|EPI_ISL_76 | ···················· | ···················· | ···················· | ···················· | ···················· | ···················· |
| AB | 4148 hCoV-19/USA/NY-Wadsworth-223831-01/2020\|EPI_ISL_76 | ···················· | ···················· | ···················· | ···················· | ···················· | ···················· |
| AB | 4149 hCoV-19/USA/NY-Wadsworth-224047-01/2020\|EPI_ISL_76 | ···················· | ·A·················· | ···················· | ···················· | ···················· | ···················· |
| AB | 4150 hCoV-19/USA/NY-Wadsworth-223742-01/2020\|EPI_ISL_76 | ···················· | ·A·················· | ···················· | ···················· | ···················· | ···················· |
| AB | 4151 hCoV-19/USA/NY-Wadsworth-224559-01/2020\|EPI_ISL_76 | ···················· | ···················· | ···················· | ···················· | ···················· | ···················· |
| AB | 4152 hCoV-19/USA/NY-Wadsworth-290315-01/2020\|EPI_ISL_76 | ···················· | ···················· | ···················· | ···················· | ···················· | ···················· |
| AB | 4153 hCoV-19/USA/NY-Wadsworth-290316-01/2020\|EPI_ISL_76 | ···················· | ·A·················· | ···················· | ···················· | ···················· | ···················· |
| AB | 4154 hCoV-19/USA/NY-Wadsworth-290318-01/2020\|EPI_ISL_76 | ···················· | ···················· | ···················· | ···················· | ···················· | ···················· |
| AB | 4155 hCoV-19/USA/NY-Wadsworth-290319-01/2020\|EPI_ISL_76 | ···················· | ···················· | ···················· | ···················· | ···················· | ···················· |
| AB | 4156 hCoV-19/USA/NY-Wadsworth-290320-01/2020\|EPI_ISL_76 | ···················· | ···················· | ···················· | ···················· | ···················· | ···················· |
| AB | 4157 hCoV-19/USA/NY-Wadsworth-290321-01/2020\|EPI_ISL_76 | ···················· | ·A·················· | ···················· | ···················· | ···················· | ···················· |
| AB | 4158 hCoV-19/USA/NY-Wadsworth-290324-01/2020\|EPI_ISL_76 | ···················· | ·A·················· | ···················· | ···················· | ···················· | ···················· |
| AB | 4159 hCoV-19/USA/NY-Wadsworth-290325-01/2020\|EPI_ISL_76 | ···················· | ···················· | ···················· | ···················· | ···················· | ···················· |
| AB | 4160 hCoV-19/USA/NY-Wadsworth-290326-01/2020\|EPI_ISL_76 | ···················· | ···················· | ···················· | ···················· | ···················· | ···················· |
| AB | 4161 hCoV-19/USA/NY-Wadsworth-290327-01/2020\|EPI_ISL_76 | ···················· | ···················· | ···················· | ···················· | ···················· | ···················· |
| AB | 4162 hCoV-19/USA/NY-Wadsworth-290328-01/2020\|EPI_ISL_76 | ···················· | ·A·················· | ···················· | ···················· | ···················· | ···················· |
| AB | 4163 hCoV-19/USA/NY-Wadsworth-290329-01/2020\|EPI_ISL_76 | ···················· | ···················· | ···················· | ···················· | ···················· | ···················· |
| AB | 4164 hCoV-19/USA/NY-Wadsworth-290330-01/2020\|EPI_ISL_76 | ···················· | ···················· | ···················· | ···················· | ···················· | ···················· |
| AB | 4165 hCoV-19/USA/NY-Wadsworth-290334-01/2020\|EPI_ISL_76 | ···················· | ···················· | ···················· | ···················· | ···················· | ···················· |
| AB | 4166 hCoV-19/USA/NY-Wadsworth-290336-01/2020\|EPI_ISL_76 | ···················· | ·A·················· | ···················· | ···················· | ···················· | ···················· |
| AB | 4167 hCoV-19/USA/NY-Wadsworth-290337-01/2020\|EPI_ISL_76 | ···················· | ···················· | ···················· | ···················· | ···················· | ···················· |
| AB | 4168 hCoV-19/USA/NY-Wadsworth-290340-01/2020\|EPI_ISL_76 | ···················· | ···················· | ···················· | ···················· | ···················· | ···················· |
| AB | 4169 hCoV-19/USA/NY-Wadsworth-290342-01/2020\|EPI_ISL_76 | ···················· | ···················· | ···················· | ···················· | ···················· | ···················· |
| AB | 4170 hCoV-19/USA/NY-Wadsworth-290345-01/2020\|EPI_ISL_76 | ···················· | ···················· | ···················· | ···················· | ···················· | ···················· |
| AB | 4171 hCoV-19/USA/NY-Wadsworth-290346-01/2020\|EPI_ISL_76 | ···················· | ·A·················· | ···················· | ···················· | ···················· | ···················· |
| AB | 4172 hCoV-19/USA/NY-Wadsworth-290348-01/2020\|EPI_ISL_76 | ···················· | ·A·················· | ···················· | ···················· | ···················· | ···················· |
| AB | 4173 hCoV-19/USA/NY-Wadsworth-290349-01/2020\|EPI_ISL_76 | ···················· | ···················· | ···················· | ···················· | ···················· | ···················· |
| AB | 4174 hCoV-19/USA/NY-Wadsworth-290351-01/2020\|EPI_ISL_76 | ···················· | ···················· | ···················· | ···················· | ···················· | ···················· |
| AB | 4175 hCoV-19/USA/NY-Wadsworth-290354-01/2020\|EPI_ISL_76 | ···················· | ·············A······ | ···················· | ···················· | ···················· | ···················· |
| AB | 4176 hCoV-19/USA/NY-Wadsworth-290356-01/2020\|EPI_ISL_76 | ···················· | ···················· | ···················· | ···················· | ···················· | ···················· |
| AB | 4177 hCoV-19/USA/NY-Wadsworth-290358-01/2020\|EPI_ISL_76 | ···················· | ···················· | ···················· | ···················· | ···················· | ···················· |

| | | | | | | | | |
|---|---|---|---|---|---|---|---|---|
| AB | 4178 | hCoV-19/USA/NY-Wadsworth-290359-01/2020\|EPI_ISL_76 | ···················· | ···················· | ···················· | ···················· | ···················· | ···················· |
| AB | 4179 | hCoV-19/USA/NY-Wadsworth-290361-01/2020\|EPI_ISL_76 | ···················· | ···················· | ···················· | ···················· | ···················· | ···················· |
| AB | 4180 | hCoV-19/USA/NY-Wadsworth-290362-01/2020\|EPI_ISL_76 | ···················· | ···················· | ···················· | ···················· | ···················· | ···················· |
| AB | 4181 | hCoV-19/USA/NY-Wadsworth-290365-01/2020\|EPI_ISL_76 | ···················· | ···················· | ···················· | ···················· | ···················· | ···················· |
| AB | 4182 | hCoV-19/USA/NY-Wadsworth-290366-01/2020\|EPI_ISL_76 | ···················· | ···················· | ···················· | ···················· | ···················· | ···················· |
| AB | 4183 | hCoV-19/USA/NY-Wadsworth-290367-01/2020\|EPI_ISL_76 | ···················· | ···················· | ···················· | ···················· | ···················· | ···················· |
| AB | 4184 | hCoV-19/USA/NY-Wadsworth-290368-01/2020\|EPI_ISL_76 | ···················· | ·A·················· | ···················· | ···················· | ···················· | ···················· |
| AB | 4185 | hCoV-19/USA/NY-Wadsworth-290369-01/2020\|EPI_ISL_76 | ···················· | ·A·················· | ···················· | ···················· | ···················· | ···················· |
| AB | 4186 | hCoV-19/USA/NY-Wadsworth-290370-01/2020\|EPI_ISL_76 | ···················· | ···················· | ···················· | ···················· | ···················· | ···················· |
| AB | 4187 | hCoV-19/USA/NY-Wadsworth-290373-01/2020\|EPI_ISL_76 | ···················· | ···················· | ···················· | ···················· | ···················· | ···················· |
| AB | 4188 | hCoV-19/USA/NY-Wadsworth-290374-01/2020\|EPI_ISL_76 | ···················· | ·A·················· | ···················· | ···················· | ···················· | ···················· |
| AB | 4189 | hCoV-19/USA/NY-Wadsworth-290375-01/2020\|EPI_ISL_76 | ···················· | ···················· | ···················· | ···················· | ···················· | ···················· |
| AB | 4190 | hCoV-19/USA/NY-Wadsworth-290376-01/2020\|EPI_ISL_76 | ···················· | ·A·················· | ···················· | ···················· | ···················· | ···················· |
| AB | 4191 | hCoV-19/USA/NY-Wadsworth-290377-01/2020\|EPI_ISL_76 | ···················· | ···················· | ···················· | ···················· | ···················· | ···················· |
| AB | 4192 | hCoV-19/USA/NY-Wadsworth-290378-01/2020\|EPI_ISL_76 | ···················· | ···················· | ···················· | ···················· | ···················· | ···················· |
| AB | 4193 | hCoV-19/England/205000381/2020\|EPI_ISL_766853\|2020 | ···················· | ···················· | ···················· | ···················· | ···················· | ···················· |
| AB | 4194 | hCoV-19/England/205260455/2020\|EPI_ISL_766859\|2020 | ···················· | ···················· | ···················· | ···················· | ···················· | ···················· |
| AB | 4195 | hCoV-19/Algeria/G41498-8846/2020\|EPI_ISL_766861\|20 | ···················· | ···················· | ···················· | ···················· | ···················· | ···················· |
| AB | 4196 | hCoV-19/Algeria/G42452-8847/2020\|EPI_ISL_766862\|20 | ···················· | ···················· | ···················· | ···················· | ···················· | ···················· |
| AB | 4197 | hCoV-19/Algeria/G37318-8849/2020\|EPI_ISL_766863\|20 | ···················· | ·A·················· | ···················· | ···················· | ···················· | ··A················· |
| AB | 4198 | hCoV-19/Algeria/G35155-8850/2020\|EPI_ISL_766864\|20 | ···················· | ···················· | ···················· | ···················· | ···················· | ···················· |
| AB | 4199 | hCoV-19/Algeria/G38218-8852/2020\|EPI_ISL_766865\|20 | ···················· | ···················· | ···················· | ···················· | ···················· | ···················· |
| AB | 4200 | hCoV-19/Algeria/G38882-8853/2020\|EPI_ISL_766866\|20 | ···················· | ···················· | ···················· | ···················· | ···················· | ···················· |
| AB | 4201 | hCoV-19/Algeria/G37138-8854/2020\|EPI_ISL_766867\|20 | ···················· | ···················· | ···················· | ···················· | ···················· | ···················· |
| AB | 4202 | hCoV-19/Algeria/G33030-8855/2020\|EPI_ISL_766868\|20 | ·······T············ | ···················· | ···················· | ···················· | ···················· | ···················· |
| AB | 4203 | hCoV-19/Algeria/G35014-8856/2020\|EPI_ISL_766869\|20 | ···················· | ···················· | ···················· | ···················· | ···················· | ···················· |
| AB | 4204 | hCoV-19/Algeria/G38599-8859/2020\|EPI_ISL_766871\|20 | ···················· | ···················· | ···················· | ···················· | ···················· | ···················· |
| AB | 4205 | hCoV-19/Algeria/G38029-8861/2020\|EPI_ISL_766872\|20 | ···················· | ·A·················· | ···················· | ···················· | ···················· | ···················· |
| AB | 4206 | hCoV-19/Algeria/G40009-8862/2020\|EPI_ISL_766873\|20 | ···················· | ···················· | ···················· | ···················· | ···················· | ···················· |
| AB | 4207 | hCoV-19/Algeria/G43521-8863/2020\|EPI_ISL_766874\|20 | ···················· | ···················· | ···················· | ···················· | ···················· | ···················· |
| AB | 4208 | hCoV-19/Algeria/G37308-8864/2020\|EPI_ISL_766875\|20 | ···················· | ·A·················· | ···················· | ···················· | ···················· | ···················· |
| AB | 4209 | hCoV-19/USA/WY-WPHL-20063196/2020\|EPI_ISL_766877\|2 | ···················· | ···················· | ···················· | ···················· | ···················· | ···················· |
| AB | 4210 | hCoV-19/USA/WY-WPHL-20063053/2020\|EPI_ISL_766878\|2 | ···················· | ·A·················· | ···················· | ···················· | ···················· | ···················· |
| AB | 4211 | hCoV-19/USA/WY-WPHL-20150412/2020\|EPI_ISL_766879\|2 | ···················· | ···················· | ···················· | ···················· | ···················· | ···················· |
| AB | 4212 | hCoV-19/USA/WY-WPHL-20149631/2020\|EPI_ISL_766880\|2 | ···················· | ·····A·············· | ···················· | ···················· | ···················· | ···················· |
| AB | 4213 | hCoV-19/USA/WY-WPHL-20149594/2020\|EPI_ISL_766881\|2 | ···················· | ·A·················· | ···················· | ···················· | ···················· | ···················· |
| AB | 4214 | hCoV-19/USA/WY-WPHL-20095145/2020\|EPI_ISL_766882\|2 | ···················· | ···················· | ···················· | ···················· | ···················· | ···················· |
| AB | 4215 | hCoV-19/Wales/ALDP-C22FC3/2020\|EPI_ISL_767250\|2020 | ···················· | ···················· | ···················· | ···················· | ···················· | ···················· |
| AB | 4216 | hCoV-19/Wales/ALDP-C3955E/2020\|EPI_ISL_767251\|2020 | ···················· | ···················· | ···················· | ···················· | ···················· | ···················· |
| AB | 4217 | hCoV-19/Wales/ALDP-C3959A/2020\|EPI_ISL_767252\|2020 | ···················· | ···················· | ···················· | ···················· | ···················· | ···················· |
| AB | 4218 | hCoV-19/Wales/ALDP-C3A71B/2020\|EPI_ISL_767255\|2020 | ···················· | ···················· | ···················· | ···················· | ···················· | ···················· |
| AB | 4219 | hCoV-19/Wales/ALDP-C3A8BE/2020\|EPI_ISL_767256\|2020 | ···················· | ···················· | ···················· | ···················· | ···················· | ···················· |
| AB | 4220 | hCoV-19/Wales/ALDP-C3A58A/2020\|EPI_ISL_767257\|2020 | ···················· | ···················· | ···················· | ···················· | ···················· | ···················· |
| AB | 4221 | hCoV-19/Wales/ALDP-C3A7DF/2020\|EPI_ISL_767258\|2020 | ···················· | ···················· | ···················· | ···················· | ···················· | ···················· |
| AB | 4222 | hCoV-19/Wales/ALDP-C3A6B4/2020\|EPI_ISL_767259\|2020 | ···················· | ···················· | ···················· | ···················· | ···················· | ···················· |
| AB | 4223 | hCoV-19/England/CAMC-C709D7/2020\|EPI_ISL_769080\|20 | ···················· | ···················· | ···················· | ···················· | ···················· | ···················· |
| AB | 4224 | hCoV-19/England/CAMC-C720FE/2020\|EPI_ISL_769082\|20 | ···················· | ·A·················· | ···················· | ···················· | ···················· | ···················· |
| AB | 4225 | hCoV-19/England/CAMC-C6FE54/2020\|EPI_ISL_769091\|20 | ···················· | ···················· | ···················· | ···················· | ···················· | ···················· |
| AB | 4226 | hCoV-19/England/CAMC-C71C37/2020\|EPI_ISL_769093\|20 | ···················· | ·A·················· | ···················· | ···················· | ···················· | ··A················· |
| AB | 4227 | hCoV-19/England/CAMC-C71B2B/2020\|EPI_ISL_769094\|20 | ···················· | ···················· | ···················· | ···················· | ···················· | ···················· |
| AB | 4228 | hCoV-19/England/CAMC-C73804/2020\|EPI_ISL_769097\|20 | ···················· | ···················· | ···················· | ···················· | ···················· | ···················· |
| AB | 4229 | hCoV-19/USA/WY-WPHL-20148996/2020\|EPI_ISL_770069\|2 | ···················· | ·A·················· | ···················· | ···················· | ···················· | ···················· |
| AB | 4230 | hCoV-19/USA/WY-WPHL-20149500/2020\|EPI_ISL_770071\|2 | ···················· | ···················· | ···················· | ···················· | ···················· | ···················· |
| AB | 4231 | hCoV-19/USA/WY-WPHL-20063191/2020\|EPI_ISL_770072\|2 | ···················· | ··C················· | ···················· | ···················· | ···················· | ···················· |
| AB | 4232 | hCoV-19/USA/WY-WPHL-20063192/2020\|EPI_ISL_770073\|2 | ···················· | ··C················· | ···················· | ···················· | ···················· | ···················· |
| AB | 4233 | hCoV-19/USA/WY-WPHL-20063197/2020\|EPI_ISL_770074\|2 | ···················· | ···················· | ···················· | ···················· | ···················· | ···················· |
| AB | 4234 | hCoV-19/USA/WY-WPHL-20063232/2020\|EPI_ISL_770075\|2 | ···················· | ··C················· | ···················· | ···················· | ···················· | ···················· |
| AB | 4235 | hCoV-19/USA/WY-WPHL-20063233/2020\|EPI_ISL_770076\|2 | ···················· | ··C················· | ···················· | ···················· | ···················· | ···················· |
| AB | 4236 | hCoV-19/USA/WY-WPHL-20063393/2020\|EPI_ISL_770077\|2 | ···················· | ···················· | ···················· | ···················· | ···················· | ···················· |
| AB | 4237 | hCoV-19/USA/WY-WPHL-20063394/2020\|EPI_ISL_770078\|2 | ···················· | ···················· | ···················· | ···················· | ···················· | ···················· |
| AB | 4238 | hCoV-19/USA/WY-WPHL-20094742/2020\|EPI_ISL_770079\|2 | ···················· | ···················· | ···················· | ···················· | ···················· | ···················· |
| AB | 4239 | hCoV-19/USA/WY-WPHL-20094746/2020\|EPI_ISL_770080\|2 | ···················· | ···················· | ···················· | ···················· | ···················· | ···················· |
| AB | 4240 | hCoV-19/USA/WY-WPHL-20095100/2020\|EPI_ISL_770081\|2 | ···················· | ···················· | ···················· | ···················· | ···················· | ···················· |
| AB | 4241 | hCoV-19/USA/WY-WPHL-20095106/2020\|EPI_ISL_770082\|2 | ···················· | ·A·················· | ···················· | ···················· | ···················· | ···················· |
| AB | 4242 | hCoV-19/USA/WY-WPHL-20095205/2020\|EPI_ISL_770083\|2 | ···················· | ···················· | ···················· | ···················· | ···················· | ···················· |
| AB | 4243 | hCoV-19/USA/WY-WPHL-20095287/2020\|EPI_ISL_770084\|2 | ···················· | ···················· | ···················· | ···················· | ···················· | ···················· |
| AB | 4244 | hCoV-19/USA/WY-WPHL-20095809/2020\|EPI_ISL_770085\|2 | ···················· | ···················· | ···················· | ···················· | ···················· | ···················· |
| AB | 4245 | hCoV-19/USA/WY-WPHL-20148235/2020\|EPI_ISL_770086\|2 | ···················· | ···················· | ···················· | ···················· | ···················· | ···················· |
| AB | 4246 | hCoV-19/USA/WY-WPHL-20148257/2020\|EPI_ISL_770087\|2 | ···················· | ···················· | ···················· | ···················· | ···················· | ···················· |
| AB | 4247 | hCoV-19/USA/WY-WPHL-20148369/2020\|EPI_ISL_770088\|2 | ···················· | ···················· | ···················· | ···················· | ···················· | ···················· |
| AB | 4248 | hCoV-19/USA/WY-WPHL-20148596/2020\|EPI_ISL_770089\|2 | ···················· | ·A·················· | ···················· | ···················· | ···················· | ···················· |
| AB | 4249 | hCoV-19/USA/WY-WPHL-20148626/2020\|EPI_ISL_770090\|2 | ···················· | ············A······· | ···········T········ | ···················· | ···················· | ···················· |
| AB | 4250 | hCoV-19/USA/WY-WPHL-20148634/2020\|EPI_ISL_770091\|2 | ···················· | ···················· | ···················· | ···················· | ···················· | ···················· |
| AB | 4251 | hCoV-19/USA/WY-WPHL-20148935/2020\|EPI_ISL_770092\|2 | ···················· | ···················· | ···················· | ···················· | ···················· | ···················· |
| AB | 4252 | hCoV-19/USA/WY-WPHL-20148999/2020\|EPI_ISL_770093\|2 | ···················· | ···················· | ···················· | ···················· | ···················· | ···················· |
| AB | 4253 | hCoV-19/USA/WY-WPHL-20149274/2020\|EPI_ISL_770094\|2 | ···················· | ···················· | ···················· | ···················· | ···················· | ···················· |
| AB | 4254 | hCoV-19/USA/WY-WPHL-20149286/2020\|EPI_ISL_770095\|2 | ···················· | ···················· | ···················· | ···················· | ···················· | ···················· |
| AB | 4255 | hCoV-19/USA/WY-WPHL-20149418/2020\|EPI_ISL_770096\|2 | ···················· | ···················· | ···················· | ···················· | ···················· | ···················· |
| AB | 4256 | hCoV-19/USA/WY-WPHL-20149462/2020\|EPI_ISL_770097\|2 | ···················· | ···················· | ···················· | ···················· | ···················· | ···················· |
| AB | 4257 | hCoV-19/USA/WY-WPHL-20149480/2020\|EPI_ISL_770098\|2 | ···················· | ···················· | ···················· | ···················· | ···················· | ···················· |

| | | | | | | | |
|---|---|---|---|---|---|---|---|
| AB | 4258 hCoV-19/USA/WY-WPHL-20149523/2020\|EPI_ISL_770099\|2 | ···················· | ·A·················· | ···················· | ···················· | ···················· | ···················· |
| AB | 4259 hCoV-19/Luxembourg/LNS6178589/2020\|EPI_ISL_770882\| | ···················· | ···················· | ···················· | ···················· | ···················· | ···················· |
| AB | 4260 hCoV-19/Luxembourg/LNS7015346/2020\|EPI_ISL_770888\| | ···················· | ···················· | ···················· | ···················· | ···················· | ···················· |
| AB | 4261 hCoV-19/Luxembourg/LNS5809570/2020\|EPI_ISL_770889\| | ···················· | ···················· | ···················· | ···················· | ···················· | ···················· |
| AB | 4262 hCoV-19/Luxembourg/LNS9718765/2020\|EPI_ISL_770890\| | ···················· | ···················· | ···················· | ···················· | ···················· | ···················· |
| AB | 4263 hCoV-19/Luxembourg/LNS1652515/2020\|EPI_ISL_770891\| | ···················· | ···················· | ···················· | ···················· | ···················· | ···················· |
| AB | 4264 hCoV-19/Luxembourg/LNS7998697/2020\|EPI_ISL_770892\| | ···················· | ···················· | ···················· | ···················· | ···················· | ···················· |
| AB | 4265 hCoV-19/Germany/HH-hpi-p1850/2020\|EPI_ISL_776200\|2 | ···················· | ···················· | ···················· | ···················· | ···················· | ···················· |
| AB | 4266 hCoV-19/Germany/HH-hpi-p1851/2020\|EPI_ISL_776201\|2 | ···················· | ···················· | ···················· | ···················· | ···················· | ···················· |
| AB | 4267 hCoV-19/Germany/HH-hpi-p1852/2020\|EPI_ISL_776202\|2 | ···················· | ···················· | ···················· | ···················· | ···················· | ···················· |
| AB | 4268 hCoV-19/Germany/HH-hpi-p1854/2020\|EPI_ISL_776203\|2 | ···················· | ···················· | ···················· | ···················· | ···················· | ···················· |
| AB | 4269 hCoV-19/Germany/HH-hpi-p1855/2020\|EPI_ISL_776204\|2 | ···················· | ···················· | ···················· | ···················· | ···················· | ···················· |
| AB | 4270 hCoV-19/Germany/HH-hpi-p1856/2020\|EPI_ISL_776205\|2 | ···················· | ···················· | ···················· | ···················· | ···················· | ···················· |
| AB | 4271 hCoV-19/Germany/HH-hpi-p1919/2020\|EPI_ISL_776250\|2 | ···················· | ···················· | ···················· | ···················· | ···················· | ···················· |
| AB | 4272 hCoV-19/Germany/HH-hpi-p1920/2020\|EPI_ISL_776251\|2 | ···················· | ···················· | ···················· | ···················· | ···················· | ···················· |
| AB | 4273 hCoV-19/Germany/HH-hpi-p1921/2020\|EPI_ISL_776252\|2 | ···················· | ···················· | ···················· | ···················· | ···················· | ···················· |
| AB | 4274 hCoV-19/Germany/HH-hpi-p1922/2020\|EPI_ISL_776253\|2 | ···················· | ···················· | ···················· | ···················· | ···················· | ···················· |
| AB | 4275 hCoV-19/Germany/HH-hpi-p1923/2020\|EPI_ISL_776254\|2 | ···················· | ···················· | ···················· | ···················· | ···················· | ···················· |
| AB | 4276 hCoV-19/Germany/HH-hpi-p1925/2020\|EPI_ISL_776255\|2 | ···················· | ···················· | ···················· | ···················· | ···················· | ···················· |
| AB | 4277 hCoV-19/Germany/HH-hpi-p1926/2020\|EPI_ISL_776256\|2 | ···················· | ···················· | ···················· | ···················· | ···················· | ···················· |
| AB | 4278 hCoV-19/Germany/HH-hpi-p1928/2020\|EPI_ISL_776257\|2 | ···················· | ···················· | ···················· | ···················· | ···················· | ···················· |
| AB | 4279 hCoV-19/Germany/HH-hpi-p1929/2020\|EPI_ISL_776258\|2 | ···················· | ···················· | ···················· | ···················· | ···················· | ···················· |
| AB | 4280 hCoV-19/Germany/HH-hpi-p1930/2020\|EPI_ISL_776259\|2 | ···················· | ···················· | ···················· | ···················· | ···················· | ···················· |
| AB | 4281 hCoV-19/Germany/HH-hpi-p1932/2020\|EPI_ISL_776260\|2 | ···················· | ···················· | ···················· | ···················· | ···················· | ···················· |
| AB | 4282 hCoV-19/Germany/HH-hpi-p1933/2020\|EPI_ISL_776261\|2 | ···················· | ···················· | ···················· | ···················· | ···················· | ···················· |
| AB | 4283 hCoV-19/Germany/HH-hpi-p1934/2020\|EPI_ISL_776262\|2 | ···················· | ···················· | ···················· | ···················· | ···················· | ···················· |
| AB | 4284 hCoV-19/Germany/HH-hpi-p1935/2020\|EPI_ISL_776263\|2 | ···················· | ···················· | ···················· | ···················· | ···················· | ···················· |
| AB | 4285 hCoV-19/Germany/HH-hpi-p1936/2020\|EPI_ISL_776264\|2 | ···················· | ···················· | ···················· | ···················· | ···················· | ···················· |
| AB | 4286 hCoV-19/Germany/HH-hpi-p1937/2020\|EPI_ISL_776265\|2 | ···················· | ···················· | ···················· | ···················· | ···················· | ···················· |
| AB | 4287 hCoV-19/Germany/HH-hpi-p1938/2020\|EPI_ISL_776266\|2 | ···················· | ···················· | ···················· | ···················· | ···················· | ···················· |
| AB | 4288 hCoV-19/Germany/HH-hpi-p1940/2020\|EPI_ISL_776267\|2 | ···················· | ···················· | ···················· | ···················· | ···················· | ···················· |
| AB | 4289 hCoV-19/Germany/HH-hpi-p1941/2020\|EPI_ISL_776268\|2 | ···················· | ···················· | ···················· | ···················· | ···················· | ···················· |
| AB | 4290 hCoV-19/Germany/HH-hpi-p1942/2020\|EPI_ISL_776269\|2 | ···················· | ···················· | ···················· | ···················· | ···················· | ···················· |
| AB | 4291 hCoV-19/England/CAMC-C9862D/2020\|EPI_ISL_777501\|20 | ···················· | ···················· | ···················· | ···················· | ···················· | ···················· |
| AB | 4292 hCoV-19/England/ALDP-C8D96E/2020\|EPI_ISL_777502\|20 | ···················· | ···················· | ···················· | ···················· | ···················· | ···················· |
| AB | 4293 hCoV-19/England/QEUH-C93EEE/2020\|EPI_ISL_777504\|20 | ···················· | ···················· | ···················· | ···················· | ···················· | ···················· |
| AB | 4294 hCoV-19/England/ALDP-C8C53E/2020\|EPI_ISL_777509\|20 | ···················· | ···················· | ···················· | ···················· | ···················· | ···················· |
| AB | 4295 hCoV-19/England/CAMC-C987C0/2020\|EPI_ISL_777511\|20 | ···················· | ···················· | ···················· | ···················· | ···················· | ···················· |
| AB | 4296 hCoV-19/England/CAMC-C99431/2020\|EPI_ISL_777513\|20 | ···················· | ···················· | ···················· | ···················· | ···················· | ···················· |
| AB | 4297 hCoV-19/England/CAMC-C98687/2020\|EPI_ISL_777521\|20 | ···················· | ···················· | ···················· | ···················· | ···················· | ···················· |
| AB | 4298 hCoV-19/England/QEUH-C94111/2020\|EPI_ISL_777523\|20 | ···················· | ···················· | ···················· | ···················· | ···················· | ···················· |
| AB | 4299 hCoV-19/England/CAMC-C98D82/2020\|EPI_ISL_777528\|20 | ···················· | ···················· | ···················· | ···················· | ···················· | ···················· |
| AB | 4300 hCoV-19/England/ALDP-C8CA3F/2020\|EPI_ISL_777532\|20 | ···················· | ···················· | ···················· | ···················· | ···················· | ···················· |
| AB | 4301 hCoV-19/England/CAMC-C984AB/2020\|EPI_ISL_777581\|20 | ···················· | ···················· | ···················· | ···················· | ···················· | ···················· |
| AB | 4302 hCoV-19/England/ALDP-C8DE41/2020\|EPI_ISL_777584\|20 | ···················· | ···················· | ···················· | ···················· | ···················· | ···················· |
| AB | 4303 hCoV-19/England/ALDP-C8DE8D/2020\|EPI_ISL_777587\|20 | ···················· | ···················· | ···················· | ···················· | ···················· | ···················· |
| AB | 4304 hCoV-19/England/ALDP-C8E9E5/2020\|EPI_ISL_777593\|20 | ···················· | ···················· | ···················· | ···················· | ···················· | ···················· |
| AB | 4305 hCoV-19/Italy/CAM-TIGEM-428/2020\|EPI_ISL_778802\|20 | ···················· | ···················· | ···················· | ···················· | ···················· | ···················· |
| AB | 4306 hCoV-19/Italy/CAM-TIGEM-504/2020\|EPI_ISL_778803\|20 | ···················· | ···················· | ···················· | ···················· | ···················· | ···················· |
| AB | 4307 hCoV-19/Italy/CAM-TIGEM-507/2020\|EPI_ISL_778805\|20 | ···················· | ···················· | ···················· | ···················· | ···················· | ···················· |
| AB | 4308 hCoV-19/Italy/CAM-TIGEM-513/2020\|EPI_ISL_778806\|20 | ···················· | ···················· | ···················· | ···················· | ···················· | ···················· |
| AB | 4309 hCoV-19/Italy/CAM-TIGEM-429/2020\|EPI_ISL_778807\|20 | ···················· | ···················· | ···················· | ···················· | ···················· | ···················· |
| AB | 4310 hCoV-19/Italy/CAM-TIGEM-514/2020\|EPI_ISL_778808\|20 | ···················· | ···················· | ···················· | ···················· | ···················· | ···················· |
| AB | 4311 hCoV-19/Italy/CAM-TIGEM-515/2020\|EPI_ISL_778809\|20 | ···················· | ···················· | ···················· | ···················· | ···················· | ···················· |
| AB | 4312 hCoV-19/Italy/CAM-TIGEM-516/2020\|EPI_ISL_778810\|20 | ···················· | ···················· | ···················· | ···················· | ···················· | ···················· |
| AB | 4313 hCoV-19/Italy/CAM-TIGEM-517/2020\|EPI_ISL_778811\|20 | ···················· | ···················· | ···················· | ···················· | ···················· | ···················· |
| AB | 4314 hCoV-19/Italy/CAM-TIGEM-518/2020\|EPI_ISL_778812\|20 | ···················· | ···················· | ···················· | ···················· | ···················· | ···················· |
| AB | 4315 hCoV-19/Italy/CAM-TIGEM-519/2020\|EPI_ISL_778813\|20 | ···················· | ···················· | ···················· | ···················· | ···················· | ···················· |
| AB | 4316 hCoV-19/Italy/CAM-TIGEM-520/2020\|EPI_ISL_778814\|20 | ···················· | ···················· | ···················· | ···················· | ···················· | ···················· |
| AB | 4317 hCoV-19/Italy/CAM-TIGEM-521/2020\|EPI_ISL_778815\|20 | ···················· | ···················· | ···················· | ···················· | ···················· | ···················· |
| AB | 4318 hCoV-19/Italy/CAM-TIGEM-522/2020\|EPI_ISL_778816\|20 | ···················· | ···················· | ···················· | ···················· | ···················· | ···················· |
| AB | 4319 hCoV-19/Italy/CAM-TIGEM-523/2020\|EPI_ISL_778817\|20 | ···················· | ···················· | ···················· | ···················· | ···················· | ···················· |
| AB | 4320 hCoV-19/Ireland/D-NVRL-AIIDV1440rpd_tst/2020\|EPI_I | ···················· | ···················· | ···················· | ···················· | ···················· | ···················· |
| AB | 4321 hCoV-19/Ireland/D-NVRL-AIIDV1440v1/2020\|EPI_ISL_77 | ···················· | ···················· | ···················· | ···················· | ···················· | ···················· |
| AB | 4322 hCoV-19/Ireland/D-NVRL-AIIDV1475v2/2020\|EPI_ISL_77 | ···················· | ···················· | ···················· | ···················· | ···················· | ···················· |
| AB | 4323 hCoV-19/Ireland/un-NVRL-AIIDV1404/2020\|EPI_ISL_778 | ···················· | ···················· | ···················· | ···················· | ···················· | ···················· |
| AB | 4324 hCoV-19/Ireland/WW-NVRL-AIIDV1434v1/2020\|EPI_ISL_7 | ···················· | ···················· | ···················· | ···················· | ···················· | ···················· |
| AB | 4325 hCoV-19/Argentina/C97133/2020\|EPI_ISL_778843\|2020- | ···················· | ···················· | ···················· | ···················· | ···················· | ···················· |
| AB | 4326 hCoV-19/Canada/ON-PHL-20-03614/2020\|EPI_ISL_778845 | ···················· | ·A·················· | ···················· | ···················· | ···················· | ··A················· |
| AB | 4327 hCoV-19/Canada/ON-PHL-20-03650/2020\|EPI_ISL_778846 | ···················· | ···················· | ···················· | ···················· | ···················· | ···················· |
| AB | 4328 hCoV-19/Canada/ON-PHL-20-03657/2020\|EPI_ISL_778847 | ···················· | ···················· | ···················· | ···················· | ···T·T·············· | ···················· |
| AB | 4329 hCoV-19/Canada/ON-PHL-20-03673/2020\|EPI_ISL_778848 | ···················· | ···················· | ···················· | ···················· | ···················· | ···················· |
| AB | 4330 hCoV-19/Canada/ON-PHL-20-03674/2020\|EPI_ISL_778849 | ···················· | ···················· | ···················· | ···················· | ···················· | ···················· |
| AB | 4332 hCoV-19/Canada/ON-PHL-20-03778/2020\|EPI_ISL_778853 | ···················· | ···················· | ···················· | ···················· | ···················· | ···················· |
| AB | 4333 hCoV-19/Canada/ON-PHL-20-03822/2020\|EPI_ISL_778854 | ···················· | ···················· | ···················· | ···················· | ···················· | ···················· |
| AB | 4334 hCoV-19/USA/OH-ODH-SC35117/2020\|EPI_ISL_778856\|202 | ···················· | ···················· | ···················· | ···················· | ···················· | ···················· |
| AB | 4335 hCoV-19/USA/OH-ODH-SC35118/2020\|EPI_ISL_778857\|202 | ···················· | ···················· | ···················· | ···················· | ···················· | ···················· |
| AB | 4336 hCoV-19/USA/OH-ODH-SC35119 /2020\|EPI_ISL_778858\|20 | ···················· | ···················· | ···················· | ···················· | ···················· | ···················· |
| AB | 4337 hCoV-19/USA/OH-ODH-SC35158/2020\|EPI_ISL_778859\|202 | ···················· | ···················· | ···················· | ···················· | ···················· | ···················· |
| AB | 4338 hCoV-19/USA/OH-ODH-SC35176 /2020\|EPI_ISL_778861\|20 | ···················· | ···················· | ···················· | ···················· | ···················· | ···················· |

| | | | | | | | | |
|---|---|---|---|---|---|---|---|---|
| AB | 4339 | hCoV-19/USA/OH-ODH-SC35177/2020\|EPI_ISL_778862\|202 | ···················· | ···················· | ···················· | ···················· | ···················· | ···················· |
| AB | 4340 | hCoV-19/USA/OH-ODH-SC35203/2020\|EPI_ISL_778864\|202 | ···················· | ···················· | ···················· | ···················· | ···················· | ···················· |
| AB | 4341 | hCoV-19/USA/RI-CDC-9KXU-8439/2020\|EPI_ISL_778866\|2 | ···················· | ···················· | ···················· | ···················· | ···················· | ···················· |
| AB | 4342 | hCoV-19/Pakistan/JRCGR-KHI32/2020\|EPI_ISL_779280\|2 | ···················· | ···················· | ···················· | ···················· | ···················· | ···················· |
| AB | 4343 | hCoV-19/Pakistan/JRCGR-KHI48/2020\|EPI_ISL_779281\|2 | ···················· | ···················· | ···················· | ···················· | ···················· | ···················· |
| AB | 4344 | hCoV-19/Pakistan/JRCGR-KHI45/2020\|EPI_ISL_779282\|2 | ···················· | ···················· | ···················· | ···················· | ···················· | ···················· |
| AB | 4345 | hCoV-19/Pakistan/JRCGR-KHI37/2020\|EPI_ISL_779283\|2 | ···················· | ·A·················· | ···················· | ···················· | ···················· | ··A················· |
| AB | 4346 | hCoV-19/Pakistan/JRCGR-KHI13/2020\|EPI_ISL_779284\|2 | ···················· | ···················· | ···················· | ···················· | ···················· | ···················· |
| AB | 4347 | hCoV-19/Pakistan/JRCGR-KHI47/2020\|EPI_ISL_779285\|2 | ···················· | ·A·················· | ···················· | ···················· | ···················· | ··A················· |
| AB | 4348 | hCoV-19/Pakistan/JRCGR-KHI16/2020\|EPI_ISL_779286\|2 | ···················· | ···················· | ···················· | ···················· | ···················· | ···················· |
| AB | 4349 | hCoV-19/Pakistan/JRCGR-KHI41/2020\|EPI_ISL_779287\|2 | ···················· | ···················· | ···················· | ···················· | ···················· | ···················· |
| AB | 4350 | hCoV-19/Pakistan/JRCGR-KHI33/2020\|EPI_ISL_779288\|2 | ···················· | ···················· | ···················· | ···················· | ···················· | ···················· |
| AB | 4351 | hCoV-19/Pakistan/JRCGR-KHI42/2020\|EPI_ISL_779289\|2 | ···················· | ···················· | ···················· | ···················· | ···················· | ···················· |
| AB | 4352 | hCoV-19/Pakistan/JRCGR-KHI17/2020\|EPI_ISL_779290\|2 | ···················· | ·A·················· | ···················· | ···················· | ···················· | ··A················· |
| AB | 4353 | hCoV-19/England/QEUH-CC32F8/2020\|EPI_ISL_760700\|20 | ···················· | ···················· | ···················· | ···················· | ···················· | ···················· |
| AB | 4354 | hCoV-19/England/QEUH-CC39A8/2020\|EPI_ISL_760704\|20 | ···················· | ···················· | ···················· | ···················· | ···················· | ···················· |
| AB | 4355 | hCoV-19/England/QEUH-CC487D/2020\|EPI_ISL_760706\|20 | ···················· | ···················· | ···················· | ···················· | ···················· | ···················· |
| AB | 4356 | hCoV-19/England/ALDP-CBE524/2020\|EPI_ISL_760710\|20 | ···················· | ···················· | ···················· | ···················· | ···················· | ···················· |
| AB | 4357 | hCoV-19/England/QEUH-CC21ED/2020\|EPI_ISL_760713\|20 | ···················· | ···················· | ···················· | ···················· | ···················· | ···················· |
| AB | 4358 | hCoV-19/England/QEUH-CC2A5B/2020\|EPI_ISL_760716\|20 | ···················· | ···················· | ···················· | ···················· | ···················· | ···················· |
| AB | 4359 | hCoV-19/England/QEUH-CC2244/2020\|EPI_ISL_760717\|20 | ···················· | ···················· | ···················· | ···················· | ···················· | ···················· |
| AB | 4360 | hCoV-19/England/QEUH-CAD18E/2020\|EPI_ISL_760720\|20 | ···················· | ···················· | ···················· | ···················· | ···················· | ···················· |
| AB | 4361 | hCoV-19/England/ALDP-CBF532/2020\|EPI_ISL_760723\|20 | ···················· | ···················· | ···················· | ···················· | ···················· | ···················· |
| AB | 4362 | hCoV-19/England/ALDP-CBE06F/2020\|EPI_ISL_760724\|20 | ···················· | ···················· | ···················· | ···················· | ···················· | ···················· |
| AB | 4363 | hCoV-19/England/ALDP-CC03F8/2020\|EPI_ISL_760726\|20 | ···················· | ···················· | ···················· | ···················· | ···················· | ···················· |
| AB | 4364 | hCoV-19/England/ALDP-CC0808/2020\|EPI_ISL_760728\|20 | ···················· | ···················· | ···················· | ···················· | ···················· | ···················· |
| AB | 4365 | hCoV-19/England/QEUH-CC23E7/2020\|EPI_ISL_760729\|20 | ···················· | ···················· | ···················· | ···················· | ···················· | ···················· |
| AB | 4366 | hCoV-19/England/QEUH-CC2E31/2020\|EPI_ISL_760730\|20 | ···················· | ···················· | ···················· | ···················· | ···················· | ···················· |
| AB | 4367 | hCoV-19/England/ALDP-CC05E3/2020\|EPI_ISL_760734\|20 | ···················· | ···················· | ···················· | ···················· | ···················· | ···················· |
| AB | 4368 | hCoV-19/England/QEUH-CC3DF7/2020\|EPI_ISL_760735\|20 | ···················· | ···················· | ···················· | ···················· | ···················· | ···················· |
| AB | 4369 | hCoV-19/England/ALDP-CBE25A/2020\|EPI_ISL_760737\|20 | ···················· | ···················· | ···················· | ···················· | ···················· | ···················· |
| AB | 4370 | hCoV-19/England/ALDP-CBF5BA/2020\|EPI_ISL_760738\|20 | ···················· | ···················· | ···················· | ···················· | ···················· | ···················· |
| AB | 4371 | hCoV-19/England/QEUH-CC2921/2020\|EPI_ISL_760741\|20 | ···················· | ···················· | ···················· | ···················· | ···················· | ···················· |
| AB | 4372 | hCoV-19/England/QEUH-CC2C64/2020\|EPI_ISL_760743\|20 | ···················· | ···················· | ···················· | ···················· | ···················· | ···················· |
| AB | 4373 | hCoV-19/England/ALDP-CC0589/2020\|EPI_ISL_760745\|20 | ···················· | ···················· | ···················· | ···················· | ·····T·············· | ···················· |
| AB | 4374 | hCoV-19/England/QEUH-CC2217/2020\|EPI_ISL_760746\|20 | ···················· | ···················· | ···················· | ···················· | ···················· | ···················· |
| AB | 4375 | hCoV-19/England/QEUH-CC3DBB/2020\|EPI_ISL_760751\|20 | ···················· | ···················· | ···················· | ···················· | ···················· | ···················· |
| AB | 4376 | hCoV-19/England/ALDP-CBE472/2020\|EPI_ISL_760752\|20 | ···················· | ···················· | ···················· | ···················· | ···················· | ···················· |
| AB | 4377 | hCoV-19/England/QEUH-CC21DE/2020\|EPI_ISL_760754\|20 | ···················· | ···················· | ···················· | ···················· | ···················· | ···················· |
| AB | 4378 | hCoV-19/England/ALDP-CBDE57/2020\|EPI_ISL_760755\|20 | ···················· | ···················· | ···················· | ···················· | ·····T·············· | ···················· |
| AB | 4379 | hCoV-19/England/ALDP-CBF3CF/2020\|EPI_ISL_760759\|20 | ···················· | ···················· | ···················· | ···················· | ···················· | ···················· |
| AB | 4380 | hCoV-19/England/QEUH-CC2CEC/2020\|EPI_ISL_760760\|20 | ···················· | ···················· | ···················· | ···················· | ···················· | ···················· |
| AB | 4381 | hCoV-19/England/ALDP-CBE894/2020\|EPI_ISL_760762\|20 | ···················· | ···················· | ···················· | ···················· | ···················· | ···················· |
| AB | 4382 | hCoV-19/England/QEUH-CC337D/2020\|EPI_ISL_760764\|20 | ···················· | ·A·················· | ···················· | ···················· | ···················· | ··A················· |
| AB | 4383 | hCoV-19/England/QEUH-CC2208/2020\|EPI_ISL_760765\|20 | ···················· | ···················· | ···················· | ···················· | ···················· | ···················· |
| AB | 4384 | hCoV-19/England/QEUH-CC335F/2020\|EPI_ISL_760766\|20 | ···················· | ···················· | ···················· | ···················· | ···················· | ···················· |
| AB | 4385 | hCoV-19/England/ALDP-CC07ED/2020\|EPI_ISL_760769\|20 | ···················· | ···················· | ···················· | ···················· | ···················· | ···················· |
| AB | 4386 | hCoV-19/England/QEUH-CC33B9/2020\|EPI_ISL_760772\|20 | ···················· | ···················· | ···················· | ···················· | ···················· | ···················· |
| AB | 4387 | hCoV-19/England/QEUH-CC3D06/2020\|EPI_ISL_760777\|20 | ···················· | ·A·······A·········· | ···················· | ···················· | ···················· | ···················· |
| AB | 4388 | hCoV-19/England/QEUH-CC4813/2020\|EPI_ISL_760779\|20 | ···················· | ···················· | ···················· | ···················· | ···················· | ···················· |
| AB | 4389 | hCoV-19/Wales/ALDP-CBF040/2020\|EPI_ISL_760780\|2020 | ···················· | ···················· | ···················· | ···················· | ···················· | ···················· |
| AB | 4390 | hCoV-19/England/QEUH-CAE85B/2020\|EPI_ISL_760782\|20 | ···················· | ···················· | ···················· | ···················· | ···················· | ···················· |
| AB | 4391 | hCoV-19/Wales/OXON-F41514/2020\|EPI_ISL_764243\|2020 | ···················· | ···················· | ···················· | ···················· | ···················· | ···················· |
| AB | 4393 | hCoV-19/Wales/PHWC-4A6035/2020\|EPI_ISL_764245\|2020 | ···················· | ···················· | ···················· | ···················· | ···················· | ···················· |
| AB | 4394 | hCoV-19/Wales/PHWC-4A4750/2020\|EPI_ISL_764246\|2020 | ···················· | ···················· | ···················· | ···················· | ···················· | ···················· |
| AB | 4395 | hCoV-19/Wales/PHWC-4A6114/2020\|EPI_ISL_764247\|2020 | ···················· | ···················· | ···················· | ···················· | ···················· | ···················· |
| AB | 4396 | hCoV-19/Wales/PHWC-4A6633/2020\|EPI_ISL_764248\|2020 | ···················· | ···················· | ···················· | ···················· | ···················· | ···················· |
| AB | 4397 | hCoV-19/Wales/PHWC-4A679A/2020\|EPI_ISL_764249\|2020 | ···················· | ···················· | ···················· | ···················· | ···················· | ···················· |
| AB | 4398 | hCoV-19/Wales/PHWC-4A391F/2020\|EPI_ISL_764251\|2020 | ···················· | ···················· | ···················· | ···················· | ···················· | ···················· |
| AB | 4399 | hCoV-19/Wales/PHWC-4A392E/2020\|EPI_ISL_764252\|2020 | ···················· | ···················· | ···················· | ···················· | ···················· | ···················· |
| AB | 4400 | hCoV-19/Wales/PHWC-4A393D/2020\|EPI_ISL_764253\|2020 | ···················· | ···················· | ···················· | ···················· | ···················· | ···················· |
| AB | 4401 | hCoV-19/Wales/PHWC-4A418F/2020\|EPI_ISL_764254\|2020 | ···················· | ···················· | ···················· | ···················· | ···················· | ···················· |
| AB | 4403 | hCoV-19/Wales/PHWC-4A58A7/2020\|EPI_ISL_764256\|2020 | ···················· | ···················· | ···················· | ···················· | ···················· | ···················· |
| AB | 4404 | hCoV-19/Wales/PHWC-4A5A29/2020\|EPI_ISL_764257\|2020 | ···················· | ···················· | ···················· | ···················· | ···················· | ···················· |
| AB | 4405 | hCoV-19/Wales/PHWC-4A5FEE/2020\|EPI_ISL_764258\|2020 | ···················· | ···················· | ···················· | ···················· | ···················· | ···················· |
| AB | 4406 | hCoV-19/Wales/PHWC-4A6017/2020\|EPI_ISL_764259\|2020 | ···················· | ···················· | ···················· | ···················· | ···················· | ···················· |
| AB | 4407 | hCoV-19/Wales/PHWC-4A6712/2020\|EPI_ISL_764260\|2020 | ···················· | ···················· | ···················· | ···················· | ···················· | ···················· |
| AB | 4408 | hCoV-19/England/CAMB-1BCA9B/2020\|EPI_ISL_764291\|20 | ···················· | ···················· | ···················· | ···················· | ···················· | ···················· |
| AB | 4409 | hCoV-19/England/CAMB-1BCB98/2020\|EPI_ISL_764296\|20 | ···················· | ···················· | ···················· | ···················· | ·····T·············· | ···················· |
| AB | 4410 | hCoV-19/England/CAMB-1BCC0E/2020\|EPI_ISL_764297\|20 | ···················· | ···················· | ···················· | ···················· | ·····T·············· | ···················· |
| AB | 4411 | hCoV-19/England/CAMB-1BCC95/2020\|EPI_ISL_764298\|20 | ···················· | ···················· | ···················· | ···················· | ···················· | ···················· |
| AB | 4412 | hCoV-19/England/CAMB-1BCCB3/2020\|EPI_ISL_764299\|20 | ···················· | ···················· | ···················· | ···················· | ···················· | ···················· |
| AB | 4413 | hCoV-19/USA/NY-Wadsworth-290379-01/2020\|EPI_ISL_76 | ···················· | ···················· | ···················· | ···················· | ···················· | ···················· |
| AB | 4414 | hCoV-19/USA/NY-Wadsworth-290381-01/2020\|EPI_ISL_76 | ···················· | ·A·················· | ···················· | ···················· | ···················· | ···················· |
| AB | 4415 | hCoV-19/USA/NY-Wadsworth-290382-01/2020\|EPI_ISL_76 | ···················· | ···················· | ···················· | ···················· | ···················· | ···················· |
| AB | 4416 | hCoV-19/USA/NY-Wadsworth-290383-01/2020\|EPI_ISL_76 | ···················· | ···················· | ···················· | ···················· | ···················· | ···················· |
| AB | 4417 | hCoV-19/USA/NY-Wadsworth-290384-01/2020\|EPI_ISL_76 | ···················· | ···················· | ···················· | ···················· | ···················· | ···················· |
| AB | 4418 | hCoV-19/USA/NY-Wadsworth-290385-01/2020\|EPI_ISL_76 | ···················· | ···················· | ···················· | ···················· | ···················· | ···················· |
| AB | 4419 | hCoV-19/USA/NY-Wadsworth-290387-01/2020\|EPI_ISL_76 | ···················· | ···················· | ···················· | ···················· | ···················· | ···················· |
| AB | 4420 | hCoV-19/USA/NY-Wadsworth-290401-01/2020\|EPI_ISL_76 | ···················· | ···················· | ···················· | ···················· | ···················· | ···················· |

| | | | | | | | | |
|---|---|---|---|---|---|---|---|---|
| AB | 4421 | hCoV-19/USA/NY-Wadsworth-290402-01/2020\|EPI_ISL_76 | ···················· | ···················· | ···················· | ···················· | ···················· | ···················· |
| AB | 4422 | hCoV-19/Italy/LAZ-INMI-130/2020\|EPI_ISL_765568\|202 | ···················· | ···················· | ···················· | ···················· | ······T············· | ···················· |
| AB | 4424 | hCoV-19/Italy/LAZ-INMI-133/2020\|EPI_ISL_765571\|202 | ···················· | ···················· | ···················· | ···················· | ···················· | ···················· |
| AB | 4425 | hCoV-19/Italy/LAZ-INMI-134/2020\|EPI_ISL_765572\|202 | ···················· | ···················· | ···················· | ···················· | ···················· | ···················· |
| AB | 4426 | hCoV-19/Italy/LAZ-INMI-135/2020\|EPI_ISL_765573\|202 | ···················· | ···················· | ···················· | ···················· | ······T············· | ···················· |
| AB | 4427 | hCoV-19/USA/CO-Broad-CMU_00007/2020\|EPI_ISL_765575 | ···················· | ···················· | ···················· | ···················· | ···················· | ···················· |
| AB | 4428 | hCoV-19/USA/CO-Broad-CMU_00010/2020\|EPI_ISL_765576 | ···················· | ···················· | ···················· | ···················· | ···················· | ···················· |
| AB | 4429 | hCoV-19/USA/CO-Broad-CMU_00113/2020\|EPI_ISL_765577 | ···················· | ···················· | ···················· | ···················· | ···················· | ···················· |
| AB | 4431 | hCoV-19/USA/CO-Broad-CMU_00118/2020\|EPI_ISL_765580 | ···················· | ···················· | ···················· | ···················· | ···················· | ···················· |
| AB | 4432 | hCoV-19/USA/CO-Broad-CMU_00123/2020\|EPI_ISL_765582 | ···················· | ···················· | ···················· | ···················· | ···················· | ···················· |
| AB | 4433 | hCoV-19/USA/MA-MGH-02662/2020\|EPI_ISL_765583\|2020- | ···················· | ···················· | ···················· | ···················· | ···················· | ···················· |
| AB | 4434 | hCoV-19/USA/MA-MGH-02690/2020\|EPI_ISL_765584\|2020- | ···················· | ···················· | ···················· | ···················· | ···················· | ···················· |
| AB | 4435 | hCoV-19/USA/MA-MGH-02691/2020\|EPI_ISL_765585\|2020- | ···················· | ···················· | ···················· | ···················· | ···················· | ···················· |
| AB | 4436 | hCoV-19/USA/MA-MGH-02692/2020\|EPI_ISL_765586\|2020- | ···················· | ···················· | ···················· | ···················· | ···················· | ···················· |
| AB | 4437 | hCoV-19/USA/MA-MGH-02695/2020\|EPI_ISL_765588\|2020- | ···················· | ···················· | ···················· | ···················· | ···················· | ···················· |
| AB | 4438 | hCoV-19/USA/MA-MGH-02696/2020\|EPI_ISL_765589\|2020- | ···················· | ···················· | ···················· | ···················· | ···················· | ···················· |
| AB | 4439 | hCoV-19/USA/MA-MGH-02699/2020\|EPI_ISL_765590\|2020- | ···················· | ···················· | ···················· | ···················· | ···················· | ···················· |
| AB | 4440 | hCoV-19/USA/MA-MGH-02702/2020\|EPI_ISL_765591\|2020- | ···················· | ···················· | ···················· | ···················· | ···················· | ···················· |
| AB | 4441 | hCoV-19/USA/MA-MGH-02703/2020\|EPI_ISL_765592\|2020- | ···················· | ···················· | ···················· | ···················· | ···················· | ···················· |
| AB | 4442 | hCoV-19/USA/MA-MGH-02705/2020\|EPI_ISL_765593\|2020- | ···················· | ···················· | ···················· | ···················· | ···················· | ···················· |
| AB | 4443 | hCoV-19/USA/MA-MGH-02707/2020\|EPI_ISL_765594\|2020- | ···················· | ···················· | ···················· | ···················· | ···················· | ···················· |
| AB | 4444 | hCoV-19/USA/MA-MGH-02709/2020\|EPI_ISL_765595\|2020- | ···················· | ···················· | ···················· | ···················· | ···················· | ···················· |
| AB | 4445 | hCoV-19/USA/MA-MGH-02714/2020\|EPI_ISL_765596\|2020- | ···················· | ···················· | ···················· | ···················· | ···················· | ···················· |
| AB | 4446 | hCoV-19/USA/MA-MGH-02715/2020\|EPI_ISL_765597\|2020- | ···················· | ···················· | ···················· | ···················· | ···················· | ···················· |
| AB | 4447 | hCoV-19/USA/MA-MGH-02718/2020\|EPI_ISL_765598\|2020- | ···················· | ···················· | ···················· | ···················· | ···················· | ···················· |
| AB | 4448 | hCoV-19/USA/MA-MGH-02721/2020\|EPI_ISL_765599\|2020- | ···················· | ···················· | ···················· | ···················· | ······T············· | ···················· |
| AB | 4449 | hCoV-19/USA/NM-DOH-2020632199/2020\|EPI_ISL_766895\| | ···················· | ···················· | ···················· | ···················· | ···················· | ···············A···· |
| AB | 4450 | hCoV-19/USA/NM-DOH-2020631000/2020\|EPI_ISL_766896\| | ···················· | ·A·················· | ···················· | ···················· | ···················· | ···A················ |
| AB | 4451 | hCoV-19/USA/NM-DOH-2020631397/2020\|EPI_ISL_766899\| | ···················· | ···················· | ···················· | ···················· | ···················· | ···················· |
| AB | 4452 | hCoV-19/USA/NM-DOH-2020631482/2020\|EPI_ISL_766900\| | ···················· | ···················· | ···················· | ···················· | ···················· | ···················· |
| AB | 4453 | hCoV-19/USA/NM-DOH-2020631558/2020\|EPI_ISL_766901\| | ···················· | ···················· | ···················· | ···················· | ···················· | ···················· |
| AB | 4454 | hCoV-19/USA/NM-DOH-2020631590/2020\|EPI_ISL_766902\| | ···················· | ···················· | ···················· | ···················· | ···················· | ···················· |
| AB | 4455 | hCoV-19/USA/NM-DOH-2020631598/2020\|EPI_ISL_766903\| | ···················· | ·A·················· | ···················· | ···················· | ···················· | ···················· |
| AB | 4456 | hCoV-19/USA/NM-DOH-2020631623/2020\|EPI_ISL_766904\| | ···················· | ···················· | ···················· | ···················· | ···················· | ···················· |
| AB | 4457 | hCoV-19/USA/NM-DOH-2020631626/2020\|EPI_ISL_766905\| | ···················· | ···················· | ···················· | ···················· | ···················· | ···················· |
| AB | 4458 | hCoV-19/USA/NM-DOH-2020631642/2020\|EPI_ISL_766906\| | ···················· | ···················· | ···················· | ···················· | ···················· | ···················· |
| AB | 4459 | hCoV-19/USA/NM-DOH-2020631725/2020\|EPI_ISL_766907\| | ···················· | ···················· | ···················· | ···················· | ···················· | ···················· |
| AB | 4460 | hCoV-19/USA/NM-DOH-2020631746/2020\|EPI_ISL_766908\| | ···················· | ···················· | ···················· | ···················· | ···················· | ···················· |
| AB | 4461 | hCoV-19/USA/NM-DOH-2020631785/2020\|EPI_ISL_766909\| | ········T··········· | ·A·················· | ···················· | ···················· | ···················· | ···················· |
| AB | 4462 | hCoV-19/USA/NM-DOH-2020631840/2020\|EPI_ISL_766910\| | ···················· | ···················· | ···················· | ···················· | ···················· | ···················· |
| AB | 4463 | hCoV-19/USA/NM-DOH-2020631927/2020\|EPI_ISL_766911\| | ···················· | ···················· | ···················· | ···················· | ···················· | ···················· |
| AB | 4464 | hCoV-19/USA/NM-DOH-2020631941/2020\|EPI_ISL_766912\| | ···········A········ | ···················· | ···················· | ···················· | ···················· | ···················· |
| AB | 4465 | hCoV-19/USA/NM-DOH-2020631996/2020\|EPI_ISL_766913\| | ···················· | ·A·················· | ···················· | ···················· | ···················· | ···················· |
| AB | 4466 | hCoV-19/USA/NM-DOH-2020632004/2020\|EPI_ISL_766914\| | ···················· | ···················· | ···················· | ···················· | ···················· | ···················· |
| AB | 4467 | hCoV-19/USA/NM-DOH-2020632022/2020\|EPI_ISL_766915\| | ···················· | ···················· | ···················· | ···················· | ···················· | ···················· |
| AB | 4468 | hCoV-19/USA/NM-DOH-2020632129/2020\|EPI_ISL_766916\| | ···················· | ···················· | ···················· | ···················· | ···················· | ···················· |
| AB | 4469 | hCoV-19/USA/NM-DOH-2020632141/2020\|EPI_ISL_766917\| | ···················· | ···················· | ···················· | ···················· | ···················· | ···················· |
| AB | 4470 | hCoV-19/USA/NM-DOH-2020632156/2020\|EPI_ISL_766918\| | ···················· | ···················· | ···················· | ···················· | ···················· | ···················· |
| AB | 4471 | hCoV-19/USA/NM-DOH-2020632174/2020\|EPI_ISL_766919\| | ···················· | ·A·················· | ···················· | ···················· | ···················· | ···················· |
| AB | 4472 | hCoV-19/USA/NM-DOH-2020632249/2020\|EPI_ISL_766920\| | ···················· | ·A·················· | ···················· | ···················· | ···················· | ···················· |
| AB | 4473 | hCoV-19/USA/NM-DOH-2020632287/2020\|EPI_ISL_766921\| | ···················· | ·A·················· | ···················· | ···················· | ···················· | ···················· |
| AB | 4474 | hCoV-19/England/CAMC-C737BC/2020\|EPI_ISL_769100\|20 | ···················· | ···················· | ···················· | ···················· | ···················· | ···················· |
| AB | 4475 | hCoV-19/England/CAMC-C76E69/2020\|EPI_ISL_769101\|20 | ···················· | ···················· | ···················· | ···················· | ···················· | ···················· |
| AB | 4476 | hCoV-19/England/CAMC-C75671/2020\|EPI_ISL_769104\|20 | ···················· | ···················· | ···················· | ···················· | ···················· | ···················· |
| AB | 4477 | hCoV-19/England/CAMC-C71B76/2020\|EPI_ISL_769106\|20 | ···················· | ···················· | ···················· | ···················· | ···················· | ···················· |
| AB | 4478 | hCoV-19/England/CAMC-C721FB/2020\|EPI_ISL_769108\|20 | ···················· | ···················· | ···················· | ···················· | ···················· | ···················· |
| AB | 4479 | hCoV-19/England/CAMC-C709E6/2020\|EPI_ISL_769110\|20 | ···················· | ···················· | ···················· | ···················· | ···················· | ···················· |
| AB | 4480 | hCoV-19/England/CAMC-C6FEBE/2020\|EPI_ISL_769112\|20 | ···················· | ···················· | ···················· | ···················· | ···················· | ···················· |
| AB | 4481 | hCoV-19/England/CAMC-C71CEC/2020\|EPI_ISL_769113\|20 | ···················· | ···················· | ···················· | ···················· | ···················· | ···················· |
| AB | 4482 | hCoV-19/England/CAMC-C72085/2020\|EPI_ISL_769127\|20 | ···················· | ···················· | ···················· | ···················· | ···················· | ···················· |
| AB | 4483 | hCoV-19/England/CAMC-C71C28/2020\|EPI_ISL_769168\|20 | ···················· | ···················· | ···················· | ···················· | ···················· | ···················· |
| AB | 4484 | hCoV-19/England/CAMC-C75644/2020\|EPI_ISL_769184\|20 | ···················· | ···················· | ···················· | ···················· | ···················· | ···················· |
| AB | 4485 | hCoV-19/England/CAMC-C71D07/2020\|EPI_ISL_769185\|20 | ···················· | ···················· | ···················· | ···················· | ···················· | ···················· |
| AB | 4486 | hCoV-19/England/CAMC-C71CBF/2020\|EPI_ISL_769190\|20 | ···················· | ···················· | ···················· | ···················· | ···················· | ···················· |
| AB | 4487 | hCoV-19/England/CAMC-C72234/2020\|EPI_ISL_769192\|20 | ···················· | ···················· | ···················· | ···················· | ···················· | ···················· |
| AB | 4488 | hCoV-19/England/CAMC-C77105/2020\|EPI_ISL_769195\|20 | ········T··········· | ···················· | ···················· | ···················· | ···················· | ···················· |
| AB | 4489 | hCoV-19/England/CAMC-C771D8/2020\|EPI_ISL_769196\|20 | ···················· | ···················· | ···················· | ···················· | ···················· | ···················· |
| AB | 4490 | hCoV-19/England/CAMC-C770DB/2020\|EPI_ISL_769198\|20 | ···················· | ···················· | ···················· | ···················· | ···················· | ···················· |
| AB | 4491 | hCoV-19/England/CAMC-C71DCB/2020\|EPI_ISL_769199\|20 | ···················· | ···················· | ···················· | ···················· | ···················· | ···················· |
| AB | 4492 | hCoV-19/USA/WY-WPHL-20149696/2020\|EPI_ISL_770100\|2 | ···················· | ···················· | ···················· | ···················· | ···················· | ···················· |
| AB | 4493 | hCoV-19/USA/WY-WPHL-20152947/2020\|EPI_ISL_770112\|2 | ···················· | ···················· | ···················· | ···················· | ···················· | ···················· |
| AB | 4494 | hCoV-19/USA/WY-WPHL-20153282/2020\|EPI_ISL_770113\|2 | ···················· | ···················· | ···················· | ···················· | ···················· | ···················· |
| AB | 4495 | hCoV-19/USA/WY-WPHL-20153326/2020\|EPI_ISL_770114\|2 | ···················· | ···················· | ···················· | ···················· | ···················· | ···················· |
| AB | 4496 | hCoV-19/USA/WY-WPHL-20153670/2020\|EPI_ISL_770115\|2 | ···················· | ···················· | ···················· | ···················· | ···················· | ···················· |
| AB | 4497 | hCoV-19/Germany/HH-hpi-p1857/2020\|EPI_ISL_776206\|2 | ···················· | ···················· | ···················· | ···················· | ···················· | ···················· |
| AB | 4498 | hCoV-19/Germany/HH-hpi-p1858/2020\|EPI_ISL_776207\|2 | ···················· | ···················· | ···················· | ···················· | ···················· | ···················· |
| AB | 4499 | hCoV-19/Germany/HH-hpi-p1860/2020\|EPI_ISL_776208\|2 | ···················· | ···················· | ···················· | ···················· | ···················· | ···················· |
| AB | 4500 | hCoV-19/Germany/HH-hpi-p1863/2020\|EPI_ISL_776209\|2 | ···················· | ···················· | ···················· | ···················· | ···················· | ···················· |
| AB | 4501 | hCoV-19/Germany/HH-hpi-p1864/2020\|EPI_ISL_776210\|2 | ···················· | ···················· | ···················· | ···················· | ···················· | ···················· |
| AB | 4502 | hCoV-19/Germany/HH-hpi-p1865/2020\|EPI_ISL_776211\|2 | ···················· | ···················· | ···················· | ···················· | ···················· | ···················· |

| | | | | | | | | |
|---|---|---|---|---|---|---|---|---|
| AB | 4503 | hCoV-19/Germany/HH-hpi-p1866/2020\|EPI_ISL_776212\|2 | ···················· | ···················· | ···················· | ···················· | ···················· | ···················· |
| AB | 4504 | hCoV-19/Germany/HH-hpi-p1867/2020\|EPI_ISL_776213\|2 | ···················· | ···················· | ···················· | ···················· | ···················· | ···················· |
| AB | 4505 | hCoV-19/Germany/HH-hpi-p1868/2020\|EPI_ISL_776214\|2 | ···················· | ···················· | ···················· | ···················· | ···················· | ···················· |
| AB | 4506 | hCoV-19/Germany/HH-hpi-p1869/2020\|EPI_ISL_776215\|2 | ···················· | ···················· | ···················· | ···················· | ···················· | ···················· |
| AB | 4507 | hCoV-19/Germany/HH-hpi-p1871/2020\|EPI_ISL_776216\|2 | ···················· | ···················· | ···················· | ···················· | ···················· | ···················· |
| AB | 4508 | hCoV-19/Germany/HH-hpi-p1872/2020\|EPI_ISL_776217\|2 | ···················· | ···················· | ···················· | ···················· | ···················· | ···················· |
| AB | 4509 | hCoV-19/Germany/HH-hpi-p1873/2020\|EPI_ISL_776218\|2 | ···················· | ···················· | ···················· | ···················· | ···················· | ···················· |
| AB | 4510 | hCoV-19/Germany/HH-hpi-p1874/2020\|EPI_ISL_776219\|2 | ···················· | ···················· | ···················· | ···················· | ···················· | ···················· |
| AB | 4511 | hCoV-19/Germany/HH-hpi-p1875/2020\|EPI_ISL_776220\|2 | ···················· | ···················· | ···················· | ···················· | ···················· | ···················· |
| AB | 4512 | hCoV-19/Germany/HH-hpi-p1876/2020\|EPI_ISL_776221\|2 | ···················· | ···················· | ···················· | ···················· | ···················· | ···················· |
| AB | 4513 | hCoV-19/Germany/HH-hpi-p1877/2020\|EPI_ISL_776222\|2 | ···················· | ···················· | ···················· | ···················· | ···················· | ···················· |
| AB | 4514 | hCoV-19/Germany/HH-hpi-p1879/2020\|EPI_ISL_776223\|2 | ···················· | ···················· | ···················· | ···················· | ···················· | ···················· |
| AB | 4515 | hCoV-19/Germany/HH-hpi-p1880/2020\|EPI_ISL_776224\|2 | ···················· | ···················· | ···················· | ···················· | ···················· | ···················· |
| AB | 4516 | hCoV-19/Germany/HH-hpi-p1881/2020\|EPI_ISL_776225\|2 | ···················· | ···················· | ···················· | ···················· | ···················· | ···················· |
| AB | 4517 | hCoV-19/Germany/HH-hpi-p1882/2020\|EPI_ISL_776226\|2 | ···················· | ···················· | ···················· | ···················· | ···················· | ···················· |
| AB | 4518 | hCoV-19/Germany/HH-hpi-p1883/2020\|EPI_ISL_776227\|2 | ···················· | ···················· | ···················· | ···················· | ···················· | ···················· |
| AB | 4519 | hCoV-19/Germany/HH-hpi-p1884/2020\|EPI_ISL_776228\|2 | ···················· | ···················· | ···················· | ···················· | ···················· | ···················· |
| AB | 4520 | hCoV-19/Germany/HH-hpi-p1885/2020\|EPI_ISL_776229\|2 | ···················· | ···················· | ···················· | ···················· | ···················· | ···················· |
| AB | 4521 | hCoV-19/Germany/HH-hpi-p1887/2020\|EPI_ISL_776230\|2 | ···················· | ···················· | ···················· | ···················· | ···················· | ···················· |
| AB | 4522 | hCoV-19/Germany/HH-hpi-p1888/2020\|EPI_ISL_776231\|2 | ········T··········· | ···················· | ···················· | ···················· | ···················· | ···················· |
| AB | 4523 | hCoV-19/Germany/HH-hpi-p1891/2020\|EPI_ISL_776232\|2 | ···················· | ···················· | ···················· | ···················· | ···················· | ···················· |
| AB | 4524 | hCoV-19/Germany/HH-hpi-p1893/2020\|EPI_ISL_776233\|2 | ···················· | ···················· | ···················· | ···················· | ···················· | ···················· |
| AB | 4525 | hCoV-19/Germany/HH-hpi-p1894/2020\|EPI_ISL_776234\|2 | ···················· | ···················· | ···················· | ···················· | ···················· | ···················· |
| AB | 4526 | hCoV-19/Germany/HH-hpi-p1895/2020\|EPI_ISL_776235\|2 | ···················· | ···················· | ···················· | ···················· | ···················· | ···················· |
| AB | 4527 | hCoV-19/Germany/HH-hpi-p1896/2020\|EPI_ISL_776236\|2 | ···················· | ···················· | ···················· | ···················· | ···················· | ···················· |
| AB | 4528 | hCoV-19/Germany/HH-hpi-p1897/2020\|EPI_ISL_776237\|2 | ···················· | ···················· | ···················· | ···················· | ···················· | ···················· |
| AB | 4529 | hCoV-19/Germany/HH-hpi-p1899/2020\|EPI_ISL_776238\|2 | ···················· | ···················· | ···················· | ···················· | ···················· | ···················· |
| AB | 4530 | hCoV-19/Germany/HH-hpi-p1900/2020\|EPI_ISL_776239\|2 | ···················· | ···················· | ···················· | ···················· | ···················· | ···················· |
| AB | 4531 | hCoV-19/Germany/HH-hpi-p1901/2020\|EPI_ISL_776240\|2 | ···················· | ···················· | ···················· | ···················· | ···················· | ···················· |
| AB | 4532 | hCoV-19/Germany/HH-hpi-p1902/2020\|EPI_ISL_776241\|2 | ···················· | ·A·················· | ···················· | ···················· | ···················· | ···················· |
| AB | 4533 | hCoV-19/Germany/HH-hpi-p1905/2020\|EPI_ISL_776242\|2 | ···················· | ···················· | ···················· | ···················· | ···················· | ···················· |
| AB | 4534 | hCoV-19/Germany/HH-hpi-p1908/2020\|EPI_ISL_776243\|2 | ···················· | ···················· | ···················· | ···················· | ···················· | ···················· |
| AB | 4536 | hCoV-19/Germany/HH-hpi-p1914/2020\|EPI_ISL_776245\|2 | ···················· | ···················· | ···················· | ···················· | ···················· | ···················· |
| AB | 4537 | hCoV-19/Germany/HH-hpi-p1915/2020\|EPI_ISL_776246\|2 | ···················· | ···················· | ···················· | ···················· | ···················· | ···················· |
| AB | 4538 | hCoV-19/Germany/HH-hpi-p1916/2020\|EPI_ISL_776247\|2 | ···················· | ···················· | ···················· | ···················· | ···················· | ···················· |
| AB | 4539 | hCoV-19/Germany/HH-hpi-p1917/2020\|EPI_ISL_776248\|2 | ···················· | ···················· | ···················· | ···················· | ···················· | ···················· |
| AB | 4540 | hCoV-19/Germany/HH-hpi-p1918/2020\|EPI_ISL_776249\|2 | ···················· | ···················· | ···················· | ···················· | ···················· | ···················· |
| AB | 4541 | hCoV-19/Germany/HH-hpi-p1943/2020\|EPI_ISL_776270\|2 | ···················· | ···················· | ···················· | ···················· | ···················· | ···················· |
| AB | 4542 | hCoV-19/Germany/HH-hpi-p1944/2020\|EPI_ISL_776271\|2 | ···················· | ···················· | ···················· | ···················· | ···················· | ···················· |
| AB | 4543 | hCoV-19/Germany/HH-hpi-p1945/2020\|EPI_ISL_776272\|2 | ···················· | ···················· | ···················· | ···················· | ···················· | ···················· |
| AB | 4544 | hCoV-19/Germany/HH-hpi-p1946/2020\|EPI_ISL_776273\|2 | ···················· | ···················· | ···················· | ···················· | ···················· | ···················· |
| AB | 4545 | hCoV-19/Germany/HH-hpi-p1947/2020\|EPI_ISL_776274\|2 | ···················· | ···················· | ···················· | ···················· | ···················· | ···················· |
| AB | 4546 | hCoV-19/Germany/HH-hpi-p1948/2020\|EPI_ISL_776275\|2 | ···················· | ···················· | ···················· | ···················· | ···················· | ···················· |
| AB | 4547 | hCoV-19/Germany/HH-hpi-p1949/2020\|EPI_ISL_776276\|2 | ···················· | ···················· | ···················· | ···················· | ···················· | ···················· |
| AB | 4548 | hCoV-19/Germany/HH-hpi-p1950/2020\|EPI_ISL_776277\|2 | ···················· | ···················· | ···················· | ···················· | ···················· | ···················· |
| AB | 4549 | hCoV-19/Germany/HH-hpi-p1951/2020\|EPI_ISL_776278\|2 | ···················· | ···················· | ···················· | ···················· | ···················· | ···················· |
| AB | 4550 | hCoV-19/Germany/HH-hpi-p1952/2020\|EPI_ISL_776279\|2 | ···················· | ···················· | ···················· | ···················· | ···················· | ···················· |
| AB | 4551 | hCoV-19/Germany/HH-hpi-p1953/2020\|EPI_ISL_776280\|2 | ···················· | ···················· | ···················· | ···················· | ···················· | ···················· |
| AB | 4552 | hCoV-19/Germany/HH-hpi-p1954/2020\|EPI_ISL_776281\|2 | ···················· | ···················· | ···················· | ···················· | ···················· | ···················· |
| AB | 4553 | hCoV-19/Germany/HH-hpi-p1957/2020\|EPI_ISL_776282\|2 | ···················· | ···················· | ···················· | ···················· | ···················· | ···················· |
| AB | 4554 | hCoV-19/Germany/HH-hpi-p1958/2020\|EPI_ISL_776283\|2 | ···················· | ···················· | ···················· | ···················· | ···················· | ···················· |
| AB | 4555 | hCoV-19/Germany/HH-hpi-p1959/2020\|EPI_ISL_776284\|2 | ···················· | ···················· | ···················· | ···················· | ···················· | ···················· |
| AB | 4556 | hCoV-19/Germany/HH-hpi-p1960/2020\|EPI_ISL_776285\|2 | ···················· | ···················· | ···················· | ···················· | ···················· | ···················· |
| AB | 4557 | hCoV-19/Germany/HH-hpi-p1961/2020\|EPI_ISL_776286\|2 | ···················· | ···················· | ···················· | ···················· | ···················· | ···················· |
| AB | 4558 | hCoV-19/Germany/HH-hpi-p1962/2020\|EPI_ISL_776287\|2 | ···················· | ···················· | ···················· | ···················· | ···················· | ···················· |
| AB | 4559 | hCoV-19/Germany/HH-hpi-p1965/2020\|EPI_ISL_776288\|2 | ···················· | ···················· | ···················· | ···················· | ···················· | ···················· |
| AB | 4560 | hCoV-19/Germany/HH-hpi-p1966/2020\|EPI_ISL_776289\|2 | ···················· | ···················· | ···················· | ···················· | ···················· | ···················· |
| AB | 4561 | hCoV-19/Germany/HH-hpi-p1967/2020\|EPI_ISL_776290\|2 | ···················· | ···················· | ···················· | ···················· | ···················· | ···················· |
| AB | 4562 | hCoV-19/Germany/HH-hpi-p1968/2020\|EPI_ISL_776291\|2 | ···················· | ···················· | ···················· | ···················· | ···················· | ···················· |
| AB | 4563 | hCoV-19/Germany/HH-hpi-p1969/2020\|EPI_ISL_776292\|2 | ···················· | ···················· | ···················· | ···················· | ···················· | ···················· |
| AB | 4564 | hCoV-19/Germany/HH-hpi-p1970/2020\|EPI_ISL_776293\|2 | ···················· | ···················· | ···················· | ···················· | ···················· | ···················· |
| AB | 4565 | hCoV-19/Germany/HH-hpi-p1973/2020\|EPI_ISL_776294\|2 | ···················· | ···················· | ···················· | ···················· | ···················· | ···················· |
| AB | 4566 | hCoV-19/Germany/HH-hpi-p1974/2020\|EPI_ISL_776295\|2 | ···················· | ···················· | ···················· | ···················· | ···················· | ···················· |
| AB | 4567 | hCoV-19/Germany/HH-hpi-p1975/2020\|EPI_ISL_776296\|2 | ···················· | ···················· | ···················· | ···················· | ···················· | ···················· |
| AB | 4568 | hCoV-19/Germany/HH-hpi-p1976/2020\|EPI_ISL_776297\|2 | ···················· | ···················· | ···················· | ···················· | ···················· | ···················· |
| AB | 4569 | hCoV-19/Germany/HH-hpi-p1978/2020\|EPI_ISL_776298\|2 | ···················· | ···················· | ···················· | ···················· | ···················· | ···················· |
| AB | 4570 | hCoV-19/Germany/HH-hpi-p1981/2020\|EPI_ISL_776299\|2 | ···················· | ···················· | ···················· | ···················· | ···················· | ···················· |
| AB | 4571 | hCoV-19/England/ALDP-C8D940/2020\|EPI_ISL_777539\|20 | ···················· | ···················· | ···················· | ···················· | ···················· | ···················· |
| AB | 4572 | hCoV-19/England/CAMC-C987DF/2020\|EPI_ISL_777547\|20 | ···················· | ···················· | ···················· | ···················· | ···················· | ···················· |
| AB | 4573 | hCoV-19/England/ALDP-C8C510/2020\|EPI_ISL_777550\|20 | ···················· | ···················· | ···················· | ···················· | ······T············· | ···················· |
| AB | 4574 | hCoV-19/England/ALDP-C8D7BF/2020\|EPI_ISL_777561\|20 | ···················· | ···················· | ···················· | ···················· | ···················· | ···················· |
| AB | 4575 | hCoV-19/England/CAMC-C98AD6/2020\|EPI_ISL_777565\|20 | ···················· | ···················· | ···················· | ···················· | ···················· | ···················· |
| AB | 4576 | hCoV-19/England/QEUH-C93E0C/2020\|EPI_ISL_777569\|20 | ···················· | ···················· | ···················· | ···················· | ···················· | ···················· |
| AB | 4577 | hCoV-19/England/ALDP-C8C501/2020\|EPI_ISL_777573\|20 | ···················· | ···················· | ···················· | ···················· | ···················· | ···················· |
| AB | 4578 | hCoV-19/England/CAMC-C99747/2020\|EPI_ISL_777575\|20 | ···················· | ···················· | ···················· | ···················· | ······T············· | ···················· |
| AB | 4579 | hCoV-19/England/ALDP-C8E55A/2020\|EPI_ISL_777576\|20 | ···················· | ···················· | ···················· | ···················· | ···················· | ···················· |
| AB | 4580 | hCoV-19/England/CAMC-C9847E/2020\|EPI_ISL_777578\|20 | ···················· | ···················· | ···················· | ···················· | ···················· | ···················· |
| AB | 4581 | hCoV-19/England/QEUH-C93E75/2020\|EPI_ISL_777579\|20 | ···················· | ···················· | ···················· | ···················· | ···················· | ···················· |
| AB | 4582 | hCoV-19/USA/MD-MDH-0486/2020\|EPI_ISL_778871\|2020-1 | ···················· | ·A·················· | ···················· | ···················· | ···················· | ···················· |
| AB | 4583 | hCoV-19/USA/MD-MDH-0487/2020\|EPI_ISL_778872\|2020-1 | ···················· | ···················· | ···················· | ···················· | ···················· | ···················· |

```
AB 4584 hCoV-19/USA/MD-MDH-0492/2020|EPI_ISL_778873|2020-1 ···················· ···················· ···················· ···················· ···················· ····················
AB 4585 hCoV-19/USA/MD-MDH-0493/2020|EPI_ISL_778874|2020-1 ···················· ···················· ···················· ···················· ···················· ····················
AB 4586 hCoV-19/USA/MD-MDH-0494/2020|EPI_ISL_778875|2020-1 ···················· ···················· ···················· ···················· ···················· ····················
AB 4587 hCoV-19/USA/MD-MDH-0495/2020|EPI_ISL_778876|2020-1 ···················· ···················· ···················· ···················· ···················· ····················
AB 4588 hCoV-19/USA/MD-MDH-0496/2020|EPI_ISL_778877|2020-1 ···················· ·A·················· ···················· ···················· ···················· ····················
AB 4589 hCoV-19/USA/MD-MDH-0497/2020|EPI_ISL_778878|2020-1 ···················· ···················· ···················· ···················· ···················· ····················
AB 4590 hCoV-19/USA/MD-MDH-0498/2020|EPI_ISL_778879|2020-1 ···················· ···················· ···················· ···················· ···················· ····················
AB 4591 hCoV-19/USA/MD-MDH-0499/2020|EPI_ISL_778880|2020-1 ···················· ···················· ···················· ···················· ···················· ····················
AB 4592 hCoV-19/USA/MD-MDH-0500/2020|EPI_ISL_778881|2020-1 ···················· ···················· ···················· ···················· ···················· ····················
AB 4593 hCoV-19/USA/MD-MDH-0501/2020|EPI_ISL_778882|2020-1 ···················· ·A·················· ···················· ···················· ···················· ····················
AB 4594 hCoV-19/USA/MD-MDH-0502/2020|EPI_ISL_778883|2020-1 ···················· ···················· ···················· ···················· ···················· ····················
AB 4595 hCoV-19/USA/MD-MDH-0503/2020|EPI_ISL_778884|2020-1 ···················· ···················· ···················· ···················· ···················· ····················
AB 4596 hCoV-19/USA/MD-MDH-0504/2020|EPI_ISL_778885|2020-1 ···················· ···················· ···················· ···················· ···················· ····················
AB 4597 hCoV-19/USA/MD-MDH-0506/2020|EPI_ISL_778887|2020-1 ···················· ···················· ···················· ···················· ···················· ····················
AB 4598 hCoV-19/USA/MD-MDH-0507/2020|EPI_ISL_778888|2020-1 ···················· ···················· ···················· ···················· ···················· ····················
AB 4599 hCoV-19/USA/MD-MDH-0510/2020|EPI_ISL_778891|2020-1 ···················· ···················· ···················· ···················· ···················· ····················
AB 4600 hCoV-19/USA/MD-MDH-0511/2020|EPI_ISL_778892|2020-1 ···················· ············C······· ···················· ···················· ···················· ····················
AB 4601 hCoV-19/USA/MD-MDH-0512/2020|EPI_ISL_778893|2020-1 ···················· ············C······· ···················· ···················· ···················· ····················
AB 4602 hCoV-19/USA/MD-MDH-0513/2020|EPI_ISL_778894|2020-1 ···················· ···················· ···················· ···················· ···················· ····················
AB 4603 hCoV-19/USA/MD-MDH-0514/2020|EPI_ISL_778895|2020-1 ···················· ···················· ···················· ···················· ···················· ····················
AB 4604 hCoV-19/USA/MD-MDH-0515/2020|EPI_ISL_778896|2020-1 ···················· ·A·················· ···················· ···················· ···················· ····················
AB 4605 hCoV-19/USA/MD-MDH-0517/2020|EPI_ISL_778897|2020-1 ···················· ·A·················· ···················· ···················· ···················· ····················
AB 4606 hCoV-19/USA/MD-MDH-0518/2020|EPI_ISL_778898|2020-1 ···················· ············C······· ···················· ···················· ···················· ····················
AB 4607 hCoV-19/USA/MD-MDH-0519/2020|EPI_ISL_778899|2020-1 ···················· ············C······· ···················· ···················· ···················· ····················
AB 4608 hCoV-19/USA/MD-MDH-0522/2020|EPI_ISL_778900|2020-1 ···················· ············C······· ···················· ···················· ···················· ····················
AB 4609 hCoV-19/England/ALDP-CBF611/2020|EPI_ISL_760785|20 ···················· ···················· ···················· ···················· ···················· ····················
AB 4610 hCoV-19/England/QEUH-CC2C82/2020|EPI_ISL_760787|20 ···················· ···················· ···················· ···················· ···················· ····················
AB 4611 hCoV-19/England/QEUH-CC2235/2020|EPI_ISL_760791|20 ···················· ···················· ···················· ···················· ···················· ····················
AB 4612 hCoV-19/England/QEUH-CC2A6A/2020|EPI_ISL_760793|20 ···················· ···················· ···················· ···················· ···················· ····················
AB 4613 hCoV-19/England/ALDP-CBF3ED/2020|EPI_ISL_760799|20 ···················· ···················· ···················· ···················· ···················· ····················
AB 4614 hCoV-19/Germany/TH-RKI-NP-0072/2020|EPI_ISL_763000 ···················· ···················· ···················· ···················· ···················· ····················
AB 4615 hCoV-19/Germany/TH-RKI-NP-0062/2020|EPI_ISL_763001 ···················· ···················· ···················· ···················· ···················· ····················
AB 4616 hCoV-19/Germany/TH-RKI-NP-0054/2020|EPI_ISL_763002 ···················· ···················· ···················· ···················· ···················· ····················
AB 4617 hCoV-19/Germany/TH-RKI-NP-0048/2020|EPI_ISL_763003 ···················· ···················· ···················· ···················· ···················· ····················
AB 4618 hCoV-19/Germany/TH-RKI-NP-0043/2020|EPI_ISL_763004 ···················· ···················· ···················· ···················· ···················· ····················
AB 4619 hCoV-19/Germany/TH-RKI-NP-0022/2020|EPI_ISL_763005 ···················· ···················· ···················· ···················· ···················· ····················
AB 4620 hCoV-19/Germany/TH-RKI-NP-0021/2020|EPI_ISL_763006 ···················· ···················· ···················· ···················· ···················· ····················
AB 4621 hCoV-19/Germany/TH-RKI-NP-0012/2020|EPI_ISL_763008 ···················· ···················· ···················· ···················· ···················· ····················
AB 4623 hCoV-19/Germany/SN-RKI-NP-0065/2020|EPI_ISL_763012 ···················· ···················· ···················· ···················· ···················· ····················
AB 4624 hCoV-19/Germany/SN-RKI-NP-0063/2020|EPI_ISL_763013 ···················· ···················· ···················· ···················· ···················· ····················
AB 4625 hCoV-19/Germany/SN-RKI-NP-0042/2020|EPI_ISL_763014 ···················· ···················· ···················· ···················· ···················· ····················
AB 4626 hCoV-19/Germany/SN-RKI-NP-0034/2020|EPI_ISL_763015 ···················· ·A·················· ···················· ···················· ···················· ···A················
AB 4627 hCoV-19/England/CAMB-1BD283/2020|EPI_ISL_764304|20 ···················· ···················· ···················· ···················· ······T············· ····················
AB 4628 hCoV-19/England/CAMB-1BD47E/2020|EPI_ISL_764305|20 ···················· ···················· ···················· ···················· ···················· ····················
AB 4629 hCoV-19/England/CAMB-1BD49C/2020|EPI_ISL_764306|20 ···················· ···················· ···················· ···················· ···················· ····················
AB 4630 hCoV-19/England/CAMB-1BD4AB/2020|EPI_ISL_764307|20 ···················· ···················· ···················· ···················· ···················· ····················
AB 4631 hCoV-19/England/CAMB-1BD4C9/2020|EPI_ISL_764308|20 ···················· ···················· ···················· ···················· ···················· ····················
AB 4632 hCoV-19/England/CAMB-1BD4D8/2020|EPI_ISL_764309|20 ···················· ···················· ···················· ···················· ···················· ····················
AB 4633 hCoV-19/England/CAMB-1BD4E7/2020|EPI_ISL_764310|20 ···················· ···················· ···················· ···················· ···················· ····················
AB 4634 hCoV-19/England/CAMB-1BD502/2020|EPI_ISL_764311|20 ···················· ···················· ···················· ···················· ···················· ····················
AB 4635 hCoV-19/England/CAMB-1BD511/2020|EPI_ISL_764312|20 ···················· ···················· ···················· ···················· ···················· ····················
AB 4636 hCoV-19/England/CAMB-1BD520/2020|EPI_ISL_764313|20 ···················· ···················· ···················· ···················· ···················· ····················
AB 4637 hCoV-19/England/CAMB-1BD53F/2020|EPI_ISL_764314|20 ···················· ···················· ···················· ···················· ···················· ····················
AB 4638 hCoV-19/England/CAMB-1BD56C/2020|EPI_ISL_764315|20 ···················· ···················· ···················· ···················· ···················· ····················
AB 4639 hCoV-19/England/CAMB-1BD57B/2020|EPI_ISL_764316|20 ···················· ···················· ···················· ···················· ···················· ····················
AB 4640 hCoV-19/England/CAMB-1BD58A/2020|EPI_ISL_764317|20 ···················· ···················· ···················· ···················· ···················· ····················
AB 4641 hCoV-19/England/CAMB-1BD5A8/2020|EPI_ISL_764318|20 ···················· ···················· ···················· ···················· ···················· ····················
AB 4642 hCoV-19/England/CAMB-1BD5C6/2020|EPI_ISL_764319|20 ···················· ···················· ···················· ···················· ···················· ····················
AB 4643 hCoV-19/England/CAMB-1BD5D5/2020|EPI_ISL_764320|20 ···················· ···················· ···················· ···················· ···················· ····················
AB 4644 hCoV-19/England/CAMB-1BD5F3/2020|EPI_ISL_764321|20 ···················· ···················· ···················· ···················· ···················· ····················
AB 4645 hCoV-19/England/CAMB-1BD60F/2020|EPI_ISL_764322|20 ···················· ···················· ···················· ···················· ···················· ····················
AB 4646 hCoV-19/England/CAMB-1BD61E/2020|EPI_ISL_764323|20 ···················· ···················· ···················· ···················· ···················· ····················
AB 4647 hCoV-19/England/CAMB-1BD62D/2020|EPI_ISL_764324|20 ···················· ···················· ···················· ···················· ···················· ····················
AB 4648 hCoV-19/England/CAMB-1BD64B/2020|EPI_ISL_764325|20 ···················· ···················· ···················· ···················· ···················· ····················
AB 4649 hCoV-19/England/CAMB-1BD669/2020|EPI_ISL_764326|20 ···················· ···················· ···················· ···················· ···················· ····················
AB 4650 hCoV-19/England/CAMB-1BD678/2020|EPI_ISL_764327|20 ···················· ···················· ···················· ···················· ···················· ····················
AB 4651 hCoV-19/England/CAMB-A97514/2020|EPI_ISL_764328|20 ···················· ···················· ···················· ···················· ···················· ····················
AB 4652 hCoV-19/England/CAMB-A97CD3/2020|EPI_ISL_764329|20 ···················· ···················· ···················· ···················· ···················· ····················
AB 4653 hCoV-19/England/CAMB-A97D0D/2020|EPI_ISL_764330|20 ···················· ···················· ···················· ···················· ···················· ····················
AB 4654 hCoV-19/England/CAMB-A97E19/2020|EPI_ISL_764331|20 ···················· ···················· ···················· ···················· ······T············· ····················
AB 4655 hCoV-19/England/CAMB-A97E37/2020|EPI_ISL_764332|20 ···················· ···················· ···················· ···················· ···················· ····················
AB 4656 hCoV-19/England/CAMB-A97E46/2020|EPI_ISL_764333|20 ···················· ···················· ···················· ···················· ···················· ····················
AB 4657 hCoV-19/England/CAMB-A97E64/2020|EPI_ISL_764334|20 ···················· ···················· ···················· ···················· ···················· ····················
AB 4658 hCoV-19/England/CAMB-A980D6/2020|EPI_ISL_764335|20 ···················· ···················· ···················· ···················· ···················· ····················
AB 4659 hCoV-19/England/CAMB-A980E5/2020|EPI_ISL_764336|20 ···················· ···················· ···················· ···················· ···················· ····················
AB 4660 hCoV-19/England/CAMB-A980F4/2020|EPI_ISL_764337|20 ···················· ···················· ···················· ···················· ···················· ····················
AB 4661 hCoV-19/England/CAMB-A98100/2020|EPI_ISL_764338|20 ···················· ···················· ···················· ···················· ···················· ····················
AB 4662 hCoV-19/England/CAMB-A9811F/2020|EPI_ISL_764339|20 ···················· ···················· ···················· ···················· ···················· ····················
AB 4663 hCoV-19/England/CAMB-A9812E/2020|EPI_ISL_764340|20 ···················· ···················· ···················· ···················· ···················· ····················
AB 4664 hCoV-19/England/CAMB-A9813D/2020|EPI_ISL_764341|20 ···················· ···················· ···················· ···················· ···················· ····················
```

| | | | | | | | | |
|---|---|---|---|---|---|---|---|---|
| AB | 4665 | hCoV-19/England/CAMB-A9814C/2020|EPI_ISL_764342|20 | ···················· | ···················· | ···················· | ···················· | ···················· | ···················· |
| AB | 4666 | hCoV-19/England/CAMB-A98391/2020|EPI_ISL_764343|20 | ···················· | ···················· | ···················· | ···················· | ·····T·············· | ···················· |
| AB | 4667 | hCoV-19/England/CAMB-A98434/2020|EPI_ISL_764344|20 | ···················· | ···················· | ···················· | ···················· | ···················· | ···················· |
| AB | 4668 | hCoV-19/England/CAMB-A9849E/2020|EPI_ISL_764345|20 | ···················· | ···················· | ···················· | ···················· | ···················· | ···················· |
| AB | 4669 | hCoV-19/England/CAMB-A984BC/2020|EPI_ISL_764346|20 | ···················· | ···················· | ···················· | ···················· | ···················· | ···················· |
| AB | 4670 | hCoV-19/England/CAMB-A984DA/2020|EPI_ISL_764347|20 | ···················· | ···················· | ···················· | ···················· | ···················· | ···················· |
| AB | 4671 | hCoV-19/USA/MA-MGH-02725/2020|EPI_ISL_765600|2020- | ···················· | ···················· | ···················· | ···················· | ···················· | ···················· |
| AB | 4672 | hCoV-19/USA/MA-MGH-02728/2020|EPI_ISL_765602|2020- | ···················· | ···················· | ···················· | ···················· | ···················· | ···················· |
| AB | 4673 | hCoV-19/USA/MA-MGH-02730/2020|EPI_ISL_765603|2020- | ···················· | ···················· | ···················· | ···················· | ···················· | ···················· |
| AB | 4674 | hCoV-19/USA/MA-MGH-02731/2020|EPI_ISL_765604|2020- | ···················· | ·A·················· | ···················· | ···················· | ···················· | ···················· |
| AB | 4675 | hCoV-19/USA/MA-MGH-02732/2020|EPI_ISL_765605|2020- | ···················· | ···················· | ···················· | ···················· | ···················· | ···················· |
| AB | 4676 | hCoV-19/USA/MA-MGH-02733/2020|EPI_ISL_765606|2020- | ···················· | ·A·················· | ···················· | ···················· | ···················· | ···················· |
| AB | 4677 | hCoV-19/USA/MA-MGH-02734/2020|EPI_ISL_765607|2020- | ···················· | ···················· | ···················· | ···················· | ···················· | ···················· |
| AB | 4678 | hCoV-19/USA/MA-MGH-02737/2020|EPI_ISL_765608|2020- | ···················· | ·A·················· | ···················· | ···················· | ···················· | ···················· |
| AB | 4679 | hCoV-19/USA/MA-MGH-02739/2020|EPI_ISL_765609|2020- | ···················· | ···················· | ···················· | ···················· | ···················· | ···················· |
| AB | 4680 | hCoV-19/USA/MA-MGH-02740/2020|EPI_ISL_765610|2020- | ···················· | ···················· | ···················· | ···················· | ···················· | ···················· |
| AB | 4681 | hCoV-19/USA/MA-MGH-02746/2020|EPI_ISL_765611|2020- | ···················· | ···················· | ···················· | ···················· | ···················· | ···················· |
| AB | 4682 | hCoV-19/USA/MA-MGH-02747/2020|EPI_ISL_765612|2020- | ···················· | ·A·················· | ···················· | ···················· | ···················· | ···················· |
| AB | 4683 | hCoV-19/USA/MA-MGH-02788/2020|EPI_ISL_765636|2020- | ···················· | ·A·················· | ···················· | ···················· | ···················· | ···················· |
| AB | 4684 | hCoV-19/USA/MA-MGH-02792/2020|EPI_ISL_765637|2020- | ···················· | ···················· | ···················· | ···················· | ···················· | ···················· |
| AB | 4685 | hCoV-19/USA/MA-MGH-02800/2020|EPI_ISL_765638|2020- | ···················· | ···················· | ···················· | ···················· | ···················· | ···················· |
| AB | 4686 | hCoV-19/USA/MA-MGH-02804/2020|EPI_ISL_765639|2020- | ···················· | ···················· | ···················· | ···················· | ···················· | ···················· |
| AB | 4687 | hCoV-19/USA/MA-MGH-02806/2020|EPI_ISL_765640|2020- | ···················· | ···················· | ···················· | ···················· | ···················· | ···················· |
| AB | 4688 | hCoV-19/USA/MA-MGH-02808/2020|EPI_ISL_765641|2020- | ···················· | ·A·················· | ···················· | ···················· | ···················· | ···················· |
| AB | 4689 | hCoV-19/USA/MA-MGH-02809/2020|EPI_ISL_765642|2020- | ···················· | ···················· | ···················· | ···················· | ···················· | ···················· |
| AB | 4690 | hCoV-19/USA/MA-MGH-02811/2020|EPI_ISL_765643|2020- | ···················· | ···················· | ···················· | ···················· | ···················· | ···················· |
| AB | 4691 | hCoV-19/USA/MA-MGH-02813/2020|EPI_ISL_765644|2020- | ···················· | ···················· | ···················· | ···················· | ···················· | ···················· |
| AB | 4692 | hCoV-19/USA/MA-MGH-02814/2020|EPI_ISL_765645|2020- | ···················· | ···················· | ···················· | ···················· | ···················· | ···················· |
| AB | 4693 | hCoV-19/USA/MA-MGH-02815/2020|EPI_ISL_765646|2020- | ···················· | ···················· | ···················· | ···················· | ···················· | ···················· |
| AB | 4694 | hCoV-19/USA/MA-MGH-02816/2020|EPI_ISL_765647|2020- | ···················· | ···················· | ···················· | ···················· | ···················· | ···················· |
| AB | 4695 | hCoV-19/USA/MA-MGH-02822/2020|EPI_ISL_765648|2020- | ···················· | ···················· | ···················· | ···················· | ···················· | ···················· |
| AB | 4696 | hCoV-19/USA/MA-MGH-02823/2020|EPI_ISL_765649|2020- | ···················· | ···················· | ···················· | ···················· | ···················· | ···················· |
| AB | 4697 | hCoV-19/USA/MA-MGH-02825/2020|EPI_ISL_765650|2020- | ···················· | ···················· | ···················· | ···················· | ···················· | ···················· |
| AB | 4698 | hCoV-19/USA/MA-MGH-02826/2020|EPI_ISL_765651|2020- | ···················· | ···················· | ···················· | ···················· | ···················· | ···················· |
| AB | 4699 | hCoV-19/USA/MA-MGH-02828/2020|EPI_ISL_765652|2020- | ···················· | ···················· | ···················· | ···················· | ···················· | ···················· |
| AB | 4700 | hCoV-19/USA/MA-MGH-02829/2020|EPI_ISL_765653|2020- | ···················· | ···················· | ···················· | ···················· | ···················· | ···················· |
| AB | 4701 | hCoV-19/USA/MA-MGH-02832/2020|EPI_ISL_765654|2020- | ···················· | ···················· | ···················· | ···················· | ···················· | ···················· |
| AB | 4702 | hCoV-19/USA/MA-MGH-02835/2020|EPI_ISL_765655|2020- | ···················· | ···················· | ···················· | ···················· | ···················· | ···················· |
| AB | 4703 | hCoV-19/USA/MA-MGH-02836/2020|EPI_ISL_765656|2020- | ···················· | ···················· | ···················· | ···················· | ···················· | ···················· |
| AB | 4704 | hCoV-19/USA/MA-MGH-02838/2020|EPI_ISL_765657|2020- | ···················· | ···················· | ···················· | ···················· | ···················· | ···················· |
| AB | 4705 | hCoV-19/USA/MA-MGH-02840/2020|EPI_ISL_765658|2020- | ···················· | ···················· | ···················· | ···················· | ···················· | ···················· |
| AB | 4706 | hCoV-19/USA/MA-MGH-02842/2020|EPI_ISL_765659|2020- | ···················· | ···················· | ···················· | ···················· | ···················· | ···················· |
| AB | 4707 | hCoV-19/USA/MA-MGH-02843/2020|EPI_ISL_765660|2020- | ···················· | ···················· | ···················· | ···················· | ···················· | ···················· |
| AB | 4708 | hCoV-19/USA/MA-MGH-02947/2020|EPI_ISL_765661|2020- | ···················· | ···················· | ···················· | ···················· | ···················· | ···················· |
| AB | 4709 | hCoV-19/USA/MA-MGH-02950/2020|EPI_ISL_765662|2020- | ···················· | ···················· | ···················· | ···················· | ···················· | ···················· |
| AB | 4710 | hCoV-19/USA/MA-MGH-02951/2020|EPI_ISL_765663|2020- | ···················· | ···················· | ···················· | ···················· | ···················· | ···················· |
| AB | 4711 | hCoV-19/USA/MA-MGH-02953/2020|EPI_ISL_765664|2020- | ···················· | ········A··········· | ···················· | ···················· | ···················· | ···················· |
| AB | 4712 | hCoV-19/USA/MA-MGH-02967/2020|EPI_ISL_765665|2020- | ···················· | ···················· | ···················· | ···················· | ···················· | ···················· |
| AB | 4713 | hCoV-19/USA/MA-MGH-02970/2020|EPI_ISL_765666|2020- | ···················· | ···················· | ···················· | ···················· | ···················· | ···················· |
| AB | 4714 | hCoV-19/USA/MA-MGH-02973/2020|EPI_ISL_765667|2020- | ···················· | ···················· | ···················· | ···················· | ···················· | ···················· |
| AB | 4715 | hCoV-19/USA/MA-MGH-02975/2020|EPI_ISL_765668|2020- | ···················· | ···················· | ···················· | ···················· | ···················· | ···················· |
| AB | 4716 | hCoV-19/USA/MA-MGH-02978/2020|EPI_ISL_765669|2020- | ···················· | ···················· | ···················· | ···················· | ···················· | ···················· |
| AB | 4717 | hCoV-19/USA/MA-MGH-02979/2020|EPI_ISL_765670|2020- | ···················· | ···················· | ···················· | ···················· | ···················· | ···················· |
| AB | 4718 | hCoV-19/USA/MA-MGH-02981/2020|EPI_ISL_765671|2020- | ···················· | ···················· | ···················· | ···················· | ···················· | ···················· |
| AB | 4719 | hCoV-19/USA/MA-MGH-02990/2020|EPI_ISL_765672|2020- | ···················· | ···················· | ···················· | ···················· | ···················· | ···················· |
| AB | 4720 | hCoV-19/USA/MA-MGH-02992/2020|EPI_ISL_765673|2020- | ···················· | ···················· | ···················· | ···················· | ···················· | ···················· |
| AB | 4721 | hCoV-19/USA/MA-MGH-02995/2020|EPI_ISL_765674|2020- | ···················· | ···················· | ···················· | ···················· | ···················· | ···················· |
| AB | 4722 | hCoV-19/USA/MA-MGH-02999/2020|EPI_ISL_765675|2020- | ···················· | ···················· | ···················· | ···················· | ···················· | ···················· |
| AB | 4723 | hCoV-19/USA/MA-MGH-03001/2020|EPI_ISL_765676|2020- | ···················· | ·A·················· | ···················· | ···················· | ···················· | ···················· |
| AB | 4724 | hCoV-19/USA/MA-MGH-03004/2020|EPI_ISL_765677|2020- | ···················· | ···················· | ···················· | ···················· | ···················· | ···················· |
| AB | 4725 | hCoV-19/USA/MA-MGH-03006/2020|EPI_ISL_765678|2020- | ···················· | ···················· | ···················· | ···················· | ···················· | ···················· |
| AB | 4726 | hCoV-19/USA/NM-DOH-2020632321/2020|EPI_ISL_766922| | ···················· | ············A······· | ···················· | ···················· | ···················· | ···················· |
| AB | 4727 | hCoV-19/USA/NM-DOH-2020632323/2020|EPI_ISL_766923| | ···················· | ···················· | ···················· | ···················· | ···················· | ···················· |
| AB | 4728 | hCoV-19/USA/NV-NSHL-00317251/2020|EPI_ISL_766938|2 | ···················· | ···················· | ···················· | ···················· | ···················· | ···················· |
| AB | 4729 | hCoV-19/USA/DE-DHSS-F987025/2020|EPI_ISL_766989|20 | ···················· | ·A·················· | ···················· | ···················· | ···················· | ···················· |
| AB | 4730 | hCoV-19/USA/DE-DHSS-F1036523/2020|EPI_ISL_766990|2 | ···················· | ···················· | ··T················· | ···················· | ···················· | ···················· |
| AB | 4731 | hCoV-19/USA/DE-DHSS-F1037845/2020|EPI_ISL_766991|2 | ···················· | ·A·················· | ···················· | ···················· | ···················· | ···················· |
| AB | 4732 | hCoV-19/USA/DE-DHSS-FLW00407322/2020|EPI_ISL_76699 | ···················· | ·A·················· | ···················· | ···················· | ···················· | ···················· |
| AB | 4733 | hCoV-19/USA/DE-DHSS-F986914/2020|EPI_ISL_766993|20 | ···················· | ···················· | ···················· | ···················· | ···················· | ···················· |
| AB | 4734 | hCoV-19/USA/DE-DHSS-FLW00408437/2020|EPI_ISL_76699 | ···················· | ···················· | ···················· | ···················· | ···················· | ···················· |
| AB | 4735 | hCoV-19/USA/DE-DHSS-F1006556/2020|EPI_ISL_766995|2 | ···················· | ·A·················· | ···················· | ···················· | ···················· | ···················· |
| AB | 4736 | hCoV-19/USA/DE-DHSS-FLW00410247/2020|EPI_ISL_76699 | ···················· | ·A·················· | ···················· | ···················· | ···················· | ···················· |
| AB | 4737 | hCoV-19/USA/DE-DHSS-FLW00404994/2020|EPI_ISL_76699 | ···················· | ·A·················· | ···················· | ···················· | ···················· | ···················· |
| AB | 4738 | hCoV-19/USA/DE-DHSS-F986562/2020|EPI_ISL_766998|20 | ···················· | ···················· | ···················· | ···················· | ···················· | ···················· |
| AB | 4739 | hCoV-19/USA/DE-DHSS-F1018845/2020|EPI_ISL_766999|2 | ···················· | ·A···A·············· | ···················· | ···················· | ···················· | ···················· |
| AB | 4740 | hCoV-19/England/CAMC-C75495/2020|EPI_ISL_769120|20 | ···················· | ···················· | ···················· | ···················· | ···················· | ···················· |
| AB | 4741 | hCoV-19/England/CAMC-C749F1/2020|EPI_ISL_769121|20 | ···················· | ···················· | ···················· | ···················· | ·····T·············· | ···················· |
| AB | 4742 | hCoV-19/England/CAMC-C75538/2020|EPI_ISL_769128|20 | ···················· | ···················· | ···················· | ···················· | ···················· | ···················· |
| AB | 4743 | hCoV-19/England/CAMC-C747E8/2020|EPI_ISL_769136|20 | ···················· | ···················· | ···················· | ···················· | ···················· | ···················· |
| AB | 4744 | hCoV-19/England/CAMC-C71CCE/2020|EPI_ISL_769154|20 | ···················· | ···················· | ···················· | ···················· | ···················· | ···················· |

```
AB    4745 hCoV-19/England/CAMC-C738AA/2020|EPI_ISL_769158|20    ····················  ····················  ····················  ····················  ····················  ····················
AB    4746 hCoV-19/England/CAMC-C71FE3/2020|EPI_ISL_769159|20    ····················  ····················  ····················  ····················  ····················  ····················
AB    4747 hCoV-19/USA/WY-WPHL-20149701/2020|EPI_ISL_770101|2    ····················  ····················  ····················  ····················  ····················  ····················
AB    4748 hCoV-19/USA/WY-WPHL-20149740/2020|EPI_ISL_770102|2    ····················  ·····A··············  ····················  ····················  ····················  ····················
AB    4749 hCoV-19/USA/WY-WPHL-20149786/2020|EPI_ISL_770103|2    ····················  ····················  ····················  ····················  ····················  ····················
AB    4750 hCoV-19/USA/WY-WPHL-20149984/2020|EPI_ISL_770104|2    ····················  ····················  ····················  ····················  ····················  ····················
AB    4751 hCoV-19/USA/WY-WPHL-20150087/2020|EPI_ISL_770105|2    ····················  ····················  ····················  ····················  ····················  ····················
AB    4752 hCoV-19/USA/WY-WPHL-20150313/2020|EPI_ISL_770106|2    ····················  ····················  ····················  ····················  ····················  ····················
AB    4753 hCoV-19/USA/WY-WPHL-20150375/2020|EPI_ISL_770107|2    ····················  ····················  ····················  ····················  ····················  ····················
AB    4754 hCoV-19/USA/WY-WPHL-20150413/2020|EPI_ISL_770108|2    ····················  ····················  ····················  ····················  ····················  ····················
AB    4755 hCoV-19/USA/WY-WPHL-20152582/2020|EPI_ISL_770109|2    ····················  ····················  ····················  ····················  ····················  ····················
AB    4756 hCoV-19/USA/WY-WPHL-20152602/2020|EPI_ISL_770110|2    ····················  ····················  ····················  ····················  ····················  ····················
AB    4757 hCoV-19/USA/WY-WPHL-20152944/2020|EPI_ISL_770111|2    ····················  ····················  ····················  ····················  ····················  ····················
AB    4758 hCoV-19/USA/WY-WPHL-20153708/2020|EPI_ISL_770116|2    ····················  ····················  ····················  ····················  ····················  ····················
AB    4759 hCoV-19/USA/WY-WPHL-20153807/2020|EPI_ISL_770117|2    ····················  ·A··················  ····················  ····················  ····················  ····················
AB    4760 hCoV-19/USA/WY-WPHL-20153943/2020|EPI_ISL_770118|2    ····················  ····················  ····················  ····················  ····················  ····················
AB    4761 hCoV-19/USA/WY-WPHL-20153958/2020|EPI_ISL_770119|2    ····················  ····················  ····················  ····················  ····················  ····················
AB    4762 hCoV-19/USA/WY-WPHL-20153965/2020|EPI_ISL_770120|2    ····················  ····················  ····················  ····················  ····················  ····················
AB    4763 hCoV-19/USA/WY-WPHL-20063090/2020|EPI_ISL_770121|2    ····················  ····················  ····················  ············N·······  ····················  ····················
AB    4764 hCoV-19/USA/WY-WPHL-20061952/2020|EPI_ISL_770124|2    ····················  ····················  ····················  ····················  ····················  ····················
AB    4765 hCoV-19/Germany/HH-hpi-p1982/2020|EPI_ISL_776300|2    ····················  ····················  ····················  ····················  ····················  ····················
AB    4766 hCoV-19/Germany/HH-hpi-p1983/2020|EPI_ISL_776301|2    ····················  ····················  ····················  ····················  ····················  ····················
AB    4767 hCoV-19/Germany/HH-hpi-p1984/2020|EPI_ISL_776302|2    ····················  ····················  ····················  ····················  ····················  ····················
AB    4768 hCoV-19/Germany/HH-hpi-p1985/2020|EPI_ISL_776303|2    ····················  ····················  ····················  ····················  ····················  ····················
AB    4769 hCoV-19/Germany/HH-hpi-p1986/2020|EPI_ISL_776304|2    ····················  ····················  ····················  ····················  ····················  ····················
AB    4770 hCoV-19/Germany/HH-hpi-p1987/2020|EPI_ISL_776305|2    ····················  ····················  ····················  ····················  ····················  ····················
AB    4771 hCoV-19/Germany/HH-hpi-p1988/2020|EPI_ISL_776306|2    ····················  ····················  ····················  ····················  ····················  ····················
AB    4772 hCoV-19/Germany/HH-hpi-p1989/2020|EPI_ISL_776307|2    ····················  ····················  ····················  ····················  ····················  ····················
AB    4773 hCoV-19/Germany/HH-hpi-p1990/2020|EPI_ISL_776308|2    ····················  ····················  ····················  ····················  ····················  ····················
AB    4774 hCoV-19/Germany/HH-hpi-p1991/2020|EPI_ISL_776309|2    ····················  ····················  ····················  ····················  ····················  ····················
AB    4775 hCoV-19/Germany/HH-hpi-p1992/2020|EPI_ISL_776310|2    ····················  ····················  ····················  ····················  ····················  ····················
AB    4776 hCoV-19/Germany/HH-hpi-p1994/2020|EPI_ISL_776311|2    ····················  ····················  ····················  ····················  ····················  ····················
AB    4777 hCoV-19/Germany/HH-hpi-p1995/2020|EPI_ISL_776312|2    ····················  ····················  ····················  ····················  ····················  ····················
AB    4778 hCoV-19/Germany/HH-hpi-p1996/2020|EPI_ISL_776313|2    ····················  ····················  ····················  ····················  ····················  ····················
AB    4779 hCoV-19/Germany/HH-hpi-p1997/2020|EPI_ISL_776314|2    ····················  ····················  ····················  ····················  ····················  ····················
AB    4780 hCoV-19/Germany/HH-hpi-p1998/2020|EPI_ISL_776315|2    ····················  ····················  ····················  ····················  ····················  ····················
AB    4781 hCoV-19/Germany/HH-hpi-p1999/2020|EPI_ISL_776316|2    ····················  ····················  ····················  ····················  ····················  ····················
AB    4782 hCoV-19/Germany/HH-hpi-p2000/2020|EPI_ISL_776317|2    ····················  ····················  ····················  ····················  ····················  ····················
AB    4783 hCoV-19/Germany/HH-hpi-p2001/2020|EPI_ISL_776318|2    ····················  ····················  ····················  ····················  ····················  ····················
AB    4784 hCoV-19/Germany/HH-hpi-p2009/2020|EPI_ISL_776319|2    ····················  ····················  ····················  ····················  ····················  ····················
AB    4785 hCoV-19/Germany/HH-hpi-p2012/2020|EPI_ISL_776320|2    ····················  ····················  ····················  ····················  ····················  ····················
AB    4786 hCoV-19/Germany/HH-hpi-p2013/2020|EPI_ISL_776321|2    ····················  ····················  ····················  ····················  ····················  ····················
AB    4787 hCoV-19/Germany/HH-hpi-p2014/2020|EPI_ISL_776322|2    ····················  ····················  ····················  ····················  ····················  ····················
AB    4788 hCoV-19/Germany/HH-hpi-p2015/2020|EPI_ISL_776323|2    ····················  ····················  ····················  ····················  ····················  ····················
AB    4789 hCoV-19/Germany/HH-hpi-p2018/2020|EPI_ISL_776324|2    ····················  ····················  ····················  ····················  ····················  ····················
AB    4790 hCoV-19/Germany/HH-hpi-p2021/2020|EPI_ISL_776325|2    ····················  ····················  ····················  ····················  ····················  ····················
AB    4791 hCoV-19/Germany/HH-hpi-p2022/2020|EPI_ISL_776326|2    ····················  ····················  ····················  ····················  ····················  ····················
AB    4792 hCoV-19/England/CAMC-C98E25/2020|EPI_ISL_777615|20    ····················  ····················  ····················  ····················  ····················  ····················
AB    4793 hCoV-19/England/CAMC-C98766/2020|EPI_ISL_777616|20    ····················  ····················  ····················  ····················  ····················  ····················
AB    4794 hCoV-19/England/QEUH-C93E2A/2020|EPI_ISL_777622|20    ····················  ····················  ····················  ····················  ····················  ····················
AB    4795 hCoV-19/England/QEUH-C946F4/2020|EPI_ISL_777623|20    ····················  ····················  ····················  ····················  ····················  ····················
AB    4796 hCoV-19/England/CAMC-C98405/2020|EPI_ISL_777624|20    ····················  ····················  ····················  ····················  ····················  ····················
AB    4797 hCoV-19/England/ALDP-C8E745/2020|EPI_ISL_777627|20    ····················  ····················  ····················  ····················  ······T·············  ····················
AB    4798 hCoV-19/England/QEUH-C93B05/2020|EPI_ISL_777630|20    ····················  ····················  ····················  ····················  ····················  ····················
AB    4799 hCoV-19/England/CAMC-C98933/2020|EPI_ISL_777632|20    ····················  ····················  ····················  ····················  ······T·············  ····················
AB    4800 hCoV-19/England/ALDP-C8E709/2020|EPI_ISL_777634|20    ····················  ····················  ····················  ····················  ····················  ····················
AB    4801 hCoV-19/England/ALDP-C8E763/2020|EPI_ISL_777639|20    ····················  ····················  ····················  ····················  ····················  ····················
AB    4802 hCoV-19/England/ALDP-C8E781/2020|EPI_ISL_777650|20    ····················  ····················  ····················  ····················  ······T·············  ····················
AB    4803 hCoV-19/England/QEUH-C93E48/2020|EPI_ISL_777657|20    ····················  ····················  ····················  ····················  ····················  ····················
AB    4804 hCoV-19/USA/MD-MDH-0523/2020|EPI_ISL_778901|2020-1    ····················  ····················  ····················  ····················  ····················  ····················
AB    4805 hCoV-19/USA/MD-MDH-0524/2020|EPI_ISL_778902|2020-1    ····················  ·A··················  ····················  ····················  ····················  ····················
AB    4806 hCoV-19/USA/MD-MDH-0526/2020|EPI_ISL_778903|2020-1    ····················  ····················  ····················  ····················  ····················  ····················
AB    4807 hCoV-19/USA/MD-MDH-0527/2020|EPI_ISL_778904|2020-1    ····················  ····················  ····················  ····················  ····················  ····················
AB    4808 hCoV-19/USA/MD-MDH-0528/2020|EPI_ISL_778905|2020-1    ····················  ····················  ····················  ····················  ····················  ····················
AB    4809 hCoV-19/USA/MD-MDH-0529/2020|EPI_ISL_778906|2020-1    ····················  ····················  ····················  ····················  ····················  ····················
AB    4810 hCoV-19/USA/MD-MDH-0530/2020|EPI_ISL_778907|2020-1    ····················  ·A··················  ····················  ····················  ····················  ····················
AB    4811 hCoV-19/USA/MD-MDH-0531/2020|EPI_ISL_778908|2020-1    ····················  ····················  ····················  ····················  ····················  ····················
AB    4812 hCoV-19/USA/MD-MDH-0532/2020|EPI_ISL_778909|2020-1    ····················  ·A··················  ····················  ····················  ····················  ····················
AB    4813 hCoV-19/USA/MD-MDH-0534/2020|EPI_ISL_778910|2020-1    ····················  ····················  ····················  ····················  ····················  ····················
AB    4814 hCoV-19/USA/MD-MDH-0536/2020|EPI_ISL_778912|2020-1    ····················  ············C·······  ····················  ····················  ····················  ····················
AB    4815 hCoV-19/USA/MD-MDH-0537/2020|EPI_ISL_778913|2020-1    ····················  ····················  ····················  ····················  ····················  ····················
AB    4816 hCoV-19/USA/MD-MDH-0538/2020|EPI_ISL_778914|2020-1    ····················  ····················  ····················  ····················  ····················  ····················
AB    4817 hCoV-19/USA/MD-MDH-0540/2020|EPI_ISL_778916|2020-1    ····················  ····················  ····················  ····················  ····················  ····················
AB    4818 hCoV-19/USA/MD-MDH-0541/2020|EPI_ISL_778917|2020-1    ····················  ····················  ····················  ····················  ····················  ····················
AB    4819 hCoV-19/USA/LA-EVTL1252/2020|EPI_ISL_778918|2020-1    ····················  ····················  ····················  ····················  ····················  ····················
AB    4820 hCoV-19/USA/LA-EVTL1253/2020|EPI_ISL_778919|2020-1    ····················  ····················  ····················  ····················  ····················  ····················
AB    4821 hCoV-19/USA/LA-EVTL1254/2020|EPI_ISL_778920|2020-1    ····················  ····················  ····················  ····················  ····················  ····················
AB    4822 hCoV-19/USA/LA-EVTL1255/2020|EPI_ISL_778921|2020-1    ····················  ····················  ····················  ····················  ····················  ····················
AB    4823 hCoV-19/USA/LA-EVTL1256/2020|EPI_ISL_778922|2020-1    ····················  ····················  ····················  ····················  ····················  ····················
AB    4824 hCoV-19/USA/LA-EVTL1279/2020|EPI_ISL_778945|2020-1    ····················  ·A··················  ····················  ····················  ····················  ····················
```

| | | | | | | | | |
|---|---|---|---|---|---|---|---|---|
| AB | 4825 | hCoV-19/USA/LA-EVTL1280/2020\|EPI_ISL_778946\|2020-1 | ···················· | ·A·················· | ····················· | ···················· | ···················· | ···················· |
| AB | 4826 | hCoV-19/USA/LA-EVTL1281/2020\|EPI_ISL_778947\|2020-1 | ···················· | ···················· | ····················· | ···················· | ···················· | ···················· |
| AB | 4827 | hCoV-19/USA/LA-EVTL1282/2020\|EPI_ISL_778948\|2020-1 | ···················· | ·A·················· | ····················· | ···················· | ···················· | ···················· |
| AB | 4828 | hCoV-19/USA/LA-EVTL1283/2020\|EPI_ISL_778949\|2020-1 | ···················· | ···················· | ····················· | ···················· | ···················· | ···················· |
| AB | 4829 | hCoV-19/USA/LA-EVTL1284/2020\|EPI_ISL_778950\|2020-1 | ···················· | ·A·················· | ····················· | ···················· | ···················· | ···················· |
| AB | 4830 | hCoV-19/USA/LA-EVTL1285/2020\|EPI_ISL_778951\|2020-1 | ···················· | ···················· | ····················· | ···················· | ···················· | ···················· |
| AB | 4831 | hCoV-19/USA/LA-EVTL1286/2020\|EPI_ISL_778952\|2020-1 | ···················· | ···················· | ····················· | ···················· | ···················· | ···················· |
| AB | 4832 | hCoV-19/USA/LA-EVTL1287/2020\|EPI_ISL_778953\|2020-1 | ···················· | ···················· | ····················· | ···················· | ···················· | ···················· |
| AB | 4833 | hCoV-19/USA/LA-EVTL1288/2020\|EPI_ISL_778954\|2020-1 | ···················· | ···················· | ····················· | ···················· | ···················· | ···················· |
| AB | 4834 | hCoV-19/USA/LA-EVTL1289/2020\|EPI_ISL_778955\|2020-1 | ···················· | ···················· | ····················· | ···················· | ···················· | ···················· |
| AB | 4835 | hCoV-19/USA/LA-EVTL1290/2020\|EPI_ISL_778956\|2020-1 | ···················· | ···················· | ····················· | ···················· | ···················· | ···················· |
| AB | 4836 | hCoV-19/USA/LA-EVTL1291/2020\|EPI_ISL_778957\|2020-1 | ···················· | ···················· | ····················· | ···················· | ···················· | ···················· |
| AB | 4837 | hCoV-19/USA/LA-EVTL1295/2020\|EPI_ISL_778961\|2020-1 | ···················· | ···················· | ····················· | ···················· | ···················· | ···················· |
| AB | 4838 | hCoV-19/USA/LA-EVTL1296/2020\|EPI_ISL_778962\|2020-1 | ···················· | ···················· | ····················· | ···················· | ···················· | ···················· |
| AB | 4839 | hCoV-19/USA/LA-EVTL1297/2020\|EPI_ISL_778963\|2020-1 | ···················· | ···················· | ····················· | ···················· | ···················· | ···················· |
| AB | 4840 | hCoV-19/USA/LA-EVTL1298/2020\|EPI_ISL_778964\|2020-1 | ···················· | ···················· | ····················· | ···················· | ···················· | ···················· |
| AB | 4841 | hCoV-19/USA/LA-EVTL1299/2020\|EPI_ISL_778965\|2020-1 | ···················· | ···················· | ····················· | ···················· | ···················· | ···················· |
| AB | 4842 | hCoV-19/USA/LA-EVTL1300/2020\|EPI_ISL_778966\|2020-1 | ···················· | ···················· | ····················· | ···················· | ···················· | ···················· |
| AB | 4843 | hCoV-19/USA/LA-EVTL1301/2020\|EPI_ISL_778967\|2020-1 | ···················· | ···················· | ····················· | ···················· | ···················· | ···················· |
| AB | 4844 | hCoV-19/USA/LA-EVTL1302/2020\|EPI_ISL_778968\|2020-1 | ···················· | ···················· | ····················· | ···················· | ···················· | ···················· |
| AB | 4845 | hCoV-19/USA/LA-EVTL1303/2020\|EPI_ISL_778969\|2020-1 | ···················· | ···················· | ····················· | ···················· | ···················· | ···················· |
| AB | 4846 | hCoV-19/USA/LA-EVTL1304/2020\|EPI_ISL_778970\|2020-1 | ···················· | ···················· | ····················· | ···················· | ···················· | ···················· |
| AB | 4847 | hCoV-19/USA/LA-EVTL1305/2020\|EPI_ISL_778971\|2020-1 | ···················· | ···················· | ····················· | ···················· | ···················· | ···················· |
| AB | 4848 | hCoV-19/USA/LA-EVTL1306/2020\|EPI_ISL_778972\|2020-1 | ···················· | ···················· | ····················· | ···················· | ···················· | ···················· |
| AB | 4849 | hCoV-19/USA/LA-EVTL1307/2020\|EPI_ISL_778973\|2020-1 | ···················· | ···················· | ····················· | ···················· | ···················· | ···················· |
| AB | 4850 | hCoV-19/USA/LA-EVTL1308/2020\|EPI_ISL_778974\|2020-1 | ···················· | ···················· | ····················· | ···················· | ···················· | ···················· |
| AB | 4851 | hCoV-19/USA/LA-EVTL1309/2020\|EPI_ISL_778975\|2020-1 | ···················· | ···················· | ····················· | ···················· | ···················· | ···················· |
| AB | 4852 | hCoV-19/USA/LA-EVTL1310/2020\|EPI_ISL_778976\|2020-1 | ···················· | ···················· | ····················· | ···················· | ···················· | ···················· |
| AB | 4853 | hCoV-19/USA/LA-EVTL1311/2020\|EPI_ISL_778977\|2020-1 | ···················· | ···················· | ····················· | ···················· | ···················· | ···················· |
| AB | 4854 | hCoV-19/USA/LA-EVTL1312/2020\|EPI_ISL_778978\|2020-1 | ···················· | ···················· | ····················· | ···················· | ···················· | ···················· |
| AB | 4855 | hCoV-19/USA/LA-EVTL1313/2020\|EPI_ISL_778979\|2020-1 | ···················· | ···················· | ····················· | ···················· | ···················· | ···················· |
| AB | 4856 | hCoV-19/USA/LA-EVTL1314/2020\|EPI_ISL_778980\|2020-1 | ···················· | ···················· | ····················· | ···················· | ···················· | ···················· |
| AB | 4857 | hCoV-19/USA/LA-EVTL1315/2020\|EPI_ISL_778981\|2020-1 | ···················· | ···················· | ····················· | ···················· | ···················· | ···················· |
| AB | 4858 | hCoV-19/USA/LA-EVTL1316/2020\|EPI_ISL_778982\|2020-1 | ···················· | ···················· | ····················· | ···················· | ···················· | ···················· |
| AB | 4859 | hCoV-19/USA/LA-EVTL1317/2020\|EPI_ISL_778983\|2020-1 | ···················· | ···················· | ····················· | ···················· | ···················· | ···················· |
| AB | 4860 | hCoV-19/USA/LA-EVTL1318/2020\|EPI_ISL_778984\|2020-1 | ···················· | ···················· | ····················· | ···················· | ···················· | ···················· |
| AB | 4861 | hCoV-19/USA/LA-EVTL1319/2020\|EPI_ISL_778985\|2020-1 | ···················· | ···················· | ····················· | ···················· | ···················· | ···················· |
| AB | 4862 | hCoV-19/USA/LA-EVTL1320/2020\|EPI_ISL_778986\|2020-1 | ···················· | ···················· | ····················· | ···················· | ···················· | ···················· |
| AB | 4863 | hCoV-19/USA/LA-EVTL1321/2020\|EPI_ISL_778987\|2020-1 | ···················· | ···················· | ····················· | ···················· | ···················· | ···················· |
| AB | 4864 | hCoV-19/USA/LA-EVTL1322/2020\|EPI_ISL_778988\|2020-1 | ···················· | ···················· | ····················· | ···················· | ···················· | ···················· |
| AB | 4865 | hCoV-19/Finland/24ES15B12/2020\|EPI_ISL_759809\|2020 | ···················· | ···················· | ····················· | ···················· | ···················· | ···················· |
| AB | 4866 | hCoV-19/Finland/24ES18AE9/2020\|EPI_ISL_759810\|2020 | ···················· | ·A·················· | ····················· | ···················· | ···················· | ··A················· |
| AB | 4867 | hCoV-19/Finland/24ES21A11/2020\|EPI_ISL_759811\|2020 | ···················· | ···················· | ····················· | ···················· | ···················· | ···················· |
| AB | 4868 | hCoV-19/Finland/24MS2E3/2020\|EPI_ISL_759812\|2020-0 | ···················· | ···················· | ····················· | ···················· | ···················· | ···················· |
| AB | 4869 | hCoV-19/Finland/24MS2G3/2020\|EPI_ISL_759813\|2020-0 | ···················· | ···················· | ····················· | ···················· | ···················· | ···················· |
| AB | 4870 | hCoV-19/Finland/24MS5A4/2020\|EPI_ISL_759814\|2020-0 | ···················· | ···················· | ····················· | ···················· | ···················· | ···················· |
| AB | 4871 | hCoV-19/Finland/24MS5B10/2020\|EPI_ISL_759815\|2020- | ···················· | ···················· | ····················· | ···················· | ···················· | ···················· |
| AB | 4872 | hCoV-19/Finland/24MS5E10/2020\|EPI_ISL_759816\|2020- | ···················· | ···················· | ····················· | ···················· | ···················· | ···················· |
| AB | 4873 | hCoV-19/Finland/24MS5H7/2020\|EPI_ISL_759817\|2020-0 | ···················· | ···················· | ····················· | ···················· | ···················· | ···················· |
| AB | 4874 | hCoV-19/Finland/24LS10C4/2020\|EPI_ISL_759818\|2020- | ···················· | ·A·················· | ····················· | ···················· | ···················· | ··A················· |
| AB | 4875 | hCoV-19/Finland/24LS10D12/2020\|EPI_ISL_759819\|2020 | ···················· | ·A·················· | ····················· | ···················· | ···················· | ··A················· |
| AB | 4876 | hCoV-19/Finland/24LS10H3/2020\|EPI_ISL_759820\|2020- | ···················· | ·A·················· | ····················· | ···················· | ···················· | ··A················· |
| AB | 4877 | hCoV-19/Finland/24LS1C11/2020\|EPI_ISL_759821\|2020- | ···················· | ·A·················· | ····················· | ···················· | ···················· | ··A················· |
| AB | 4878 | hCoV-19/Finland/24LS1D9/2020\|EPI_ISL_759822\|2020-1 | ···················· | ···················· | ····················· | ···················· | ···················· | ···················· |
| AB | 4879 | hCoV-19/Finland/24LS1E5/2020\|EPI_ISL_759823\|2020-1 | ···················· | ·A·················· | ····················· | ···················· | ···················· | ··A················· |
| AB | 4880 | hCoV-19/Finland/24LS2A12/2020\|EPI_ISL_759824\|2020- | ···················· | ·A·················· | ····················· | ···················· | ···················· | ··A················· |
| AB | 4881 | hCoV-19/Finland/24LS6C12/2020\|EPI_ISL_759825\|2020- | ···················· | ·A·················· | ····················· | ···················· | ···················· | ··A················· |
| AB | 4882 | hCoV-19/Finland/24LS7G11/2020\|EPI_ISL_759826\|2020- | ···················· | ·A·················· | ····················· | ···················· | ···················· | ··A················· |
| AB | 4883 | hCoV-19/Finland/24SS10H11/2020\|EPI_ISL_759827\|2020 | ···················· | ···················· | ····················· | ···················· | ···················· | ···················· |
| AB | 4884 | hCoV-19/Finland/24SS11G1/2020\|EPI_ISL_759828\|2020- | ···················· | ·A·················· | ····················· | ···················· | ···················· | ··A················· |
| AB | 4885 | hCoV-19/Finland/24SS19H9/2020\|EPI_ISL_759829\|2020- | ···················· | ·A·················· | ····················· | ···················· | ···················· | ··A················· |
| AB | 4886 | hCoV-19/Finland/24SS1E11/2020\|EPI_ISL_759830\|2020- | ···················· | ···················· | ····················· | ···················· | ···················· | ···················· |
| AB | 4887 | hCoV-19/Finland/24SS4A5/2020\|EPI_ISL_759831\|2020-0 | ···················· | ·A·················· | ····················· | ···················· | ···················· | ··A················· |
| AB | 4888 | hCoV-19/Finland/24SS4D3/2020\|EPI_ISL_759832\|2020-0 | ···················· | ·A·················· | ····················· | ···················· | ···················· | ···················· |
| AB | 4889 | hCoV-19/Finland/24SS4F3/2020\|EPI_ISL_759833\|2020-0 | ···················· | ···················· | ····················· | ···················· | ···················· | ···················· |
| AB | 4890 | hCoV-19/Finland/24SS4G1/2020\|EPI_ISL_759834\|2020-0 | ···················· | ·A·················· | ····················· | ···················· | ···················· | ··A················· |
| AB | 4891 | hCoV-19/Finland/24SS5D8/2020\|EPI_ISL_759835\|2020-0 | ···················· | ·A·················· | ····················· | ···················· | ···················· | ··A················· |
| AB | 4892 | hCoV-19/Finland/24SS5F9/2020\|EPI_ISL_759836\|2020-0 | ···················· | ·A·················· | ····················· | ···················· | ···················· | ··A················· |
| AB | 4893 | hCoV-19/Finland/24SS6bD7/2020\|EPI_ISL_759837\|2020- | ···················· | ·A·················· | ····················· | ···················· | ···················· | ··A················· |
| AB | 4894 | hCoV-19/England/MILK-CC1C38/2020\|EPI_ISL_760801\|20 | ···················· | ···················· | ····················· | ···················· | ···················· | ···················· |
| AB | 4895 | hCoV-19/England/ALDP-CBDE1B/2020\|EPI_ISL_760802\|20 | ···················· | ···················· | ····················· | ···················· | ······T············· | ···················· |
| AB | 4896 | hCoV-19/England/ALDP-CBF56F/2020\|EPI_ISL_760804\|20 | ···················· | ···················· | ····················· | ···················· | ···················· | ···················· |
| AB | 4897 | hCoV-19/England/MILK-CB1C77/2020\|EPI_ISL_760809\|20 | ···················· | ···················· | ····················· | ···················· | ···················· | ···················· |
| AB | 4898 | hCoV-19/England/MILK-CB2C76/2020\|EPI_ISL_760810\|20 | ···················· | ···················· | ····················· | ···················· | ···················· | ···················· |
| AB | 4899 | hCoV-19/England/QEUH-CADB08/2020\|EPI_ISL_760811\|20 | ···················· | ···················· | ····················· | ···················· | ···················· | ···················· |
| AB | 4900 | hCoV-19/England/ALDP-CB455B/2020\|EPI_ISL_760812\|20 | ···················· | ···················· | ····················· | ···················· | ···················· | ···················· |
| AB | 4901 | hCoV-19/England/CAMC-CB270C/2020\|EPI_ISL_760813\|20 | ···················· | ···················· | ····················· | ···················· | ···················· | ···················· |
| AB | 4902 | hCoV-19/England/ALDP-CB42DC/2020\|EPI_ISL_760817\|20 | ···················· | ···················· | ····················· | ···················· | ···················· | ···················· |
| AB | 4903 | hCoV-19/England/QEUH-CAD85C/2020\|EPI_ISL_760818\|20 | ···················· | ···················· | ····················· | ···················· | ···················· | ···················· |
| AB | 4904 | hCoV-19/Germany/SN-RKI-NP-0028/2020\|EPI_ISL_763018 | ···················· | ···················· | ····················· | ···················· | ···················· | ···················· |

AB 4905 hCoV-19/Germany/SN-RKI-NP-0024/2020|EPI_ISL_763019 ···················· ···················· ···················· ···················· ···················· ····················
AB 4906 hCoV-19/Germany/SN-RKI-NP-0013/2020|EPI_ISL_763021 ···················· ···················· ···················· ···················· ···················· ····················
AB 4907 hCoV-19/Germany/SL-RKI-NP-0046/2020|EPI_ISL_763022 ···················· ···················· ···················· ···················· ···················· ····················
AB 4908 hCoV-19/Germany/NW-RKI-NP-0030/2020|EPI_ISL_763023 ···················· ·A·················· ···················· ···················· ···················· ····················
AB 4909 hCoV-19/Germany/NW-RKI-NP-0026/2020|EPI_ISL_763025 ···················· ···················· ···················· ···················· ···················· ····················
AB 4910 hCoV-19/Germany/HE-RKI-NP-0004/2020|EPI_ISL_763035 ···················· ···················· ···················· ···················· ···················· ····················
AB 4911 hCoV-19/Germany/BY-RKI-NP-0060/2020|EPI_ISL_763036 ···················· ···················· ···················· ···················· ···················· ····················
AB 4912 hCoV-19/Germany/BB-RKI-NP-0036/2020|EPI_ISL_763060 ···················· ···················· ···················· ···················· ···················· ····················
AB 4913 hCoV-19/Germany/BB-RKI-NP-0035/2020|EPI_ISL_763061 ···················· ···················· ···················· ···················· ···················· ····················
AB 4914 hCoV-19/Georgia/Tb-171516/2020|EPI_ISL_763062|2020 ···················· ···················· ···················· ···················· ···················· ····················
AB 4915 hCoV-19/Tunisia/6895/2020|EPI_ISL_763065|2020-04-0 ···················· ···················· ···················· ···················· ···················· ····················
AB 4916 hCoV-19/Tunisia/7899/2020|EPI_ISL_763067|2020-04-0 ···················· ···················· ···················· ···················· ···················· ····················
AB 4917 hCoV-19/Italy/UMB-IZSUM-48682-1/2020|EPI_ISL_76306 ···················· ···················· ···················· ···················· ···················· ····················
AB 4918 hCoV-19/Italy/UMB-IZSUM-48682-9/2020|EPI_ISL_76307 ···················· ···················· ···················· ···················· ···················· ····················
AB 4919 hCoV-19/Italy/UMB-IZSUM-48682-26/2020|EPI_ISL_7630 ···················· ···················· ···················· ···················· ···················· ····················
AB 4920 hCoV-19/Italy/UMB-IZSUM-48682-18/2020|EPI_ISL_7630 ···················· ···················· ···················· ···················· ···················· ····················
AB 4921 hCoV-19/USA/OH-ODH-SC35001/2020|EPI_ISL_763076|202 ···················· ···················· ···················· ···················· ···················· ····················
AB 4922 hCoV-19/Germany/TH-IIMK-CaSe-20/2020|EPI_ISL_76307 ···················· ···················· ···················· ···················· ···················· ····················
AB 4923 hCoV-19/Germany/TH-IIMK-CaSe-21/2020|EPI_ISL_76308 ···················· ···················· ···················· ···················· ···················· ····················
AB 4924 hCoV-19/Germany/TH-IIMK-CaSe-22/2020|EPI_ISL_76308 ···················· ···················· ···················· ···················· ···················· ····················
AB 4925 hCoV-19/Germany/TH-IIMK-CaSe-23/2020|EPI_ISL_76308 ···················· ···················· ···················· ···················· ···················· ····················
AB 4926 hCoV-19/Germany/TH-IIMK-CaSe-24/2020|EPI_ISL_76308 ···················· ···················· ···················· ···················· ···················· ····················
AB 4927 hCoV-19/Germany/TH-IIMK-CaSe-25/2020|EPI_ISL_76309 ···················· ···················· ···················· ···················· ···················· ····················
AB 4928 hCoV-19/Germany/TH-IIMK-CaSe-26/2020|EPI_ISL_76309 ···················· ···················· ···················· ···················· ···················· ····················
AB 4929 hCoV-19/Germany/TH-IIMK-CaSe-27/2020|EPI_ISL_76309 ···················· ···················· ···················· ···················· ···················· ····················
AB 4930 hCoV-19/Germany/TH-IIMK-CaSe-28/2020|EPI_ISL_76309 ···················· ···················· ···················· ···················· ···················· ····················
AB 4931 hCoV-19/Italy/UMB-IZSUM-48682-12/2020|EPI_ISL_7630 ···················· ···················· ···················· ···················· ···················· ····················
AB 4932 hCoV-19/Italy/UMB-IZSUM-48682-17/2020|EPI_ISL_7630 ···················· ···················· ···················· ···················· ···················· ····················
AB 4933 hCoV-19/Italy/UMB-IZSUM-48682-15/2020|EPI_ISL_7630 ···················· ···················· ···················· ···················· ···················· ····················
AB 4934 hCoV-19/Italy/UMB-IZSUM-48682-20/2020|EPI_ISL_7630 ···················· ···················· ···················· ···················· ···················· ····················
AB 4935 hCoV-19/Italy/UMB-IZSUM-48682-10/2020|EPI_ISL_7630 ···················· ···················· ···················· ···················· ···················· ····················
AB 4936 hCoV-19/Belgium/un-EMC-7/2020|EPI_ISL_763099|2020- ···················· ···················· ···················· ···················· ···················· ····················
AB 4937 hCoV-19/England/CAMB-1BCCC2/2020|EPI_ISL_764300|20 ···················· ···················· ···················· ···················· ···················· ····················
AB 4938 hCoV-19/England/CAMB-1BCD29/2020|EPI_ISL_764301|20 ···················· ···················· ···················· ···················· ···················· ····················
AB 4939 hCoV-19/England/CAMB-1BCD83/2020|EPI_ISL_764302|20 ···················· ···················· ···················· ···················· ···················· ····················
AB 4940 hCoV-19/England/CAMB-1BD0D4/2020|EPI_ISL_764303|20 ···················· ···················· ···················· ···················· ···················· ····················
AB 4941 hCoV-19/England/CAMB-A984F8/2020|EPI_ISL_764348|20 ···················· ···················· ···················· ···················· ···················· ····················
AB 4942 hCoV-19/England/CAMB-A98522/2020|EPI_ISL_764349|20 ···················· ···················· ···················· ···················· ···················· ····················
AB 4943 hCoV-19/England/CAMB-A9857D/2020|EPI_ISL_764350|20 ···················· ···················· ···················· ···················· ···················· ····················
AB 4944 hCoV-19/England/CAMB-A985B9/2020|EPI_ISL_764351|20 ···················· ···················· ···················· ···················· ···················· ····················
AB 4945 hCoV-19/England/CAMB-A985D7/2020|EPI_ISL_764352|20 ···················· ···················· ···················· ···················· ···················· ····················
AB 4946 hCoV-19/England/CAMB-A98610/2020|EPI_ISL_764353|20 ···················· ···················· ···················· ···················· ···················· ····················
AB 4947 hCoV-19/England/CAMB-A9870E/2020|EPI_ISL_764354|20 ···················· ···················· ···················· ···················· ···················· ····················
AB 4948 hCoV-19/England/CAMB-A98768/2020|EPI_ISL_764355|20 ···················· ···················· ···················· ···················· ···················· ····················
AB 4949 hCoV-19/Scotland/EDB10499/2020|EPI_ISL_764356|2020 ···················· ···················· ···················· ···················· ···················· ····················
AB 4950 hCoV-19/Scotland/EDB10793/2020|EPI_ISL_764357|2020 ···················· ···················· ···················· ···················· ···················· ····················
AB 4951 hCoV-19/Scotland/EDB11088/2020|EPI_ISL_764358|2020 ···················· ···················· ···················· ···················· ···················· ····················
AB 4952 hCoV-19/Scotland/EDB11097/2020|EPI_ISL_764360|2020 ···················· ···················· ···················· ···················· ···················· ····················
AB 4953 hCoV-19/Scotland/EDB11314/2020|EPI_ISL_764361|2020 ···················· ···················· ···················· ···················· ···················· ····················
AB 4956 hCoV-19/Northern Ireland/NIRE-234B6A/2020|EPI_ISL_ ···················· ···················· ···················· ···················· ···················· ····················
AB 4957 hCoV-19/Northern Ireland/NIRE-234B79/2020|EPI_ISL_ ···················· ···················· ···················· ···················· ···················· ····················
AB 4959 hCoV-19/Northern Ireland/NIRE-234CD0/2020|EPI_ISL_ ···················· ···················· ···················· ···················· ···················· ····················
AB 4960 hCoV-19/Northern Ireland/NIRE-234D91/2020|EPI_ISL_ ···················· ···················· ···················· ···················· ···················· ····················
AB 4961 hCoV-19/Northern Ireland/NIRE-234DFB/2020|EPI_ISL_ ···················· ···················· ···················· ···················· ···················· ····················
AB 4962 hCoV-19/Northern Ireland/NIRE-234E16/2020|EPI_ISL_ ···················· ···················· ···················· ···················· ···················· ····················
AB 4963 hCoV-19/Northern Ireland/NIRE-234E34/2020|EPI_ISL_ ···················· ···················· ···················· ···················· ···················· ····················
AB 4964 hCoV-19/Northern Ireland/NIRE-234E43/2020|EPI_ISL_ ···················· ···················· ···················· ···················· ···················· ····················
AB 4965 hCoV-19/USA/MA-MGH-02748/2020|EPI_ISL_765613|2020- ···················· ···················· ···················· ···················· ······T············· ····················
AB 4966 hCoV-19/USA/MA-MGH-02749/2020|EPI_ISL_765614|2020- ···················· ···················· ···················· ···················· ···················· ····················
AB 4967 hCoV-19/USA/MA-MGH-02750/2020|EPI_ISL_765615|2020- ···················· ···················· ···················· ···················· ···················· ····················
AB 4968 hCoV-19/USA/MA-MGH-02751/2020|EPI_ISL_765616|2020- ···················· ···················· ···················· ···················· ···················· ····················
AB 4969 hCoV-19/USA/MA-MGH-02753/2020|EPI_ISL_765617|2020- ···················· ···················· ···················· ···················· ···················· ····················
AB 4970 hCoV-19/USA/MA-MGH-02755/2020|EPI_ISL_765618|2020- ···················· ···················· ···················· ···················· ···················· ····················
AB 4971 hCoV-19/USA/MA-MGH-02757/2020|EPI_ISL_765619|2020- ···················· ···················· ···················· ···················· ···················· ····················
AB 4972 hCoV-19/USA/MA-MGH-02759/2020|EPI_ISL_765620|2020- ···················· ···················· ···················· ···················· ···················· ····················
AB 4973 hCoV-19/USA/MA-MGH-02760/2020|EPI_ISL_765621|2020- ···················· ···················· ···················· ···················· ···················· ····················
AB 4974 hCoV-19/USA/MA-MGH-02763/2020|EPI_ISL_765622|2020- ···················· ···················· ···················· ···················· ···················· ····················
AB 4975 hCoV-19/USA/MA-MGH-02765/2020|EPI_ISL_765623|2020- ···················· ···················· ···················· ···················· ···················· ····················
AB 4976 hCoV-19/USA/MA-MGH-02767/2020|EPI_ISL_765624|2020- ···················· ···················· ···················· ···················· ···················· ····················
AB 4977 hCoV-19/USA/MA-MGH-02768/2020|EPI_ISL_765625|2020- ···················· ···················· ···················· ···················· ···················· ····················
AB 4978 hCoV-19/USA/MA-MGH-02769/2020|EPI_ISL_765626|2020- ···················· ···················· ···················· ···················· ···················· ····················
AB 4979 hCoV-19/USA/MA-MGH-02771/2020|EPI_ISL_765627|2020- ···················· ···················· ···················· ···················· ···················· ····················
AB 4980 hCoV-19/USA/MA-MGH-02776/2020|EPI_ISL_765628|2020- ···················· ···················· ···················· ···················· ···················· ····················
AB 4981 hCoV-19/USA/MA-MGH-02779/2020|EPI_ISL_765629|2020- ···················· ·A·················· ···················· ···················· ···················· ····················
AB 4982 hCoV-19/USA/MA-MGH-02780/2020|EPI_ISL_765630|2020- ···················· ···················· ···················· ···················· ···················· ····················
AB 4983 hCoV-19/USA/MA-MGH-02781/2020|EPI_ISL_765631|2020- ···················· ···················· ···················· ···················· ···················· ····················
AB 4984 hCoV-19/USA/MA-MGH-02784/2020|EPI_ISL_765633|2020- ···················· ···················· ···················· ···················· ···················· ····················
AB 4985 hCoV-19/USA/MA-MGH-02785/2020|EPI_ISL_765634|2020- ···················· ···················· ···················· ···················· ···················· ····················
AB 4986 hCoV-19/USA/MA-MGH-03007/2020|EPI_ISL_765679|2020- ···················· ···················· ···················· ···················· ···················· ····················
AB 4987 hCoV-19/USA/MA-MGH-03013/2020|EPI_ISL_765680|2020- ···················· ···················· ···················· ···················· ···················· ····················

| | | | | | | | | |
|---|---|---|---|---|---|---|---|---|
| AB | 4988 hCoV-19/USA/MA-MGH-03014/2020\|EPI_ISL_765681\|2020- | ···················· | ···················· | ···················· | ···················· | ···················· | ···················· | |
| AB | 4989 hCoV-19/USA/MA-MGH-03020/2020\|EPI_ISL_765682\|2020- | ···················· | ···················· | ···········T········ | ···················· | ···················· | ···················· | |
| AB | 4990 hCoV-19/USA/MA-MGH-03021/2020\|EPI_ISL_765683\|2020- | ···················· | ···················· | ···················· | ···················· | ···················· | ···················· | |
| AB | 4991 hCoV-19/USA/MA-MGH-03028/2020\|EPI_ISL_765684\|2020- | ···················· | ···················· | ···················· | ···················· | ···················· | ···················· | |
| AB | 4992 hCoV-19/USA/MA-MGH-03029/2020\|EPI_ISL_765685\|2020- | ···················· | ···················· | ···················· | ···················· | ···················· | ···················· | |
| AB | 4993 hCoV-19/USA/MA-MGH-03031/2020\|EPI_ISL_765686\|2020- | ···················· | ···················· | ···················· | ···················· | ···················· | ···················· | |
| AB | 4994 hCoV-19/USA/MA-MGH-03033/2020\|EPI_ISL_765687\|2020- | ···················· | ···················· | ···················· | ···················· | ···················· | ···················· | |
| AB | 4995 hCoV-19/USA/MA-MGH-03034/2020\|EPI_ISL_765688\|2020- | ···················· | ···················· | ···················· | ···················· | ···················· | ···················· | |
| AB | 4996 hCoV-19/USA/MA-MGH-03040/2020\|EPI_ISL_765689\|2020- | ···················· | ···················· | ···················· | ···················· | ···················· | ···················· | |
| AB | 4997 hCoV-19/USA/MA-MGH-03042/2020\|EPI_ISL_765690\|2020- | ···················· | ···················· | ···················· | ···················· | ···················· | ···················· | |
| AB | 4998 hCoV-19/USA/MA-MGH-03043/2020\|EPI_ISL_765691\|2020- | ···················· | ···················· | ···················· | ···················· | ···················· | ···················· | |
| AB | 4999 hCoV-19/USA/MA-MGH-03047/2020\|EPI_ISL_765692\|2020- | ···················· | ···················· | ···················· | ···················· | ···················· | ···················· | |
| AB | 5000 hCoV-19/USA/MA-MGH-03075/2020\|EPI_ISL_765699\|2020- | ···················· | ···················· | ···················· | ···················· | ···················· | ···················· | |
| AB | 5001 hCoV-19/Bangladesh/DNAS_DRICM_isl_18953/2020\|EPI_I | ···················· | ···················· | ···················· | ···················· | ···················· | ···················· | |
| AB | 5002 hCoV-19/Bangladesh/DNAS_DRICM_isl_MH-25/2020\|EPI_I | ···················· | ···················· | ···················· | ···················· | ···················· | ···················· | |
| AB | 5003 hCoV-19/Bangladesh/DNAS_DRICM_isl_18944/2020\|EPI_I | ···················· | ···················· | ···················· | ···················· | ···················· | ···················· | |
| AB | 5004 hCoV-19/Bangladesh/DNAS_DRICM_isl_MLS-9/2020\|EPI_I | ···················· | ···················· | ···················· | ···················· | ···················· | ···················· | |
| AB | 5005 hCoV-19/Bangladesh/DNAS_DRICM_isl_18964/2020\|EPI_I | ···················· | ···················· | ···················· | ···················· | ···················· | ···················· | |
| AB | 5006 hCoV-19/Bangladesh/DNAS_DRICM_isl_Al-52/2020\|EPI_I | ···················· | ···················· | ···················· | ···················· | ···················· | ···················· | |
| AB | 5007 hCoV-19/Bangladesh/DNAS_DRICM_isl_S-1/2020\|EPI_ISL | ···················· | ···················· | ···················· | ···················· | ···················· | ···················· | |
| AB | 5008 hCoV-19/Bangladesh/DNAS_DRICM_isl_SM-14/2020\|EPI_I | ···················· | ···················· | ···················· | ···················· | ···················· | ···················· | |
| AB | 5009 hCoV-19/Bangladesh/DNAS_DRICM_isl_Razia/2020\|EPI_I | ···················· | ···················· | ···················· | ···················· | ···················· | ···············A···· | |
| AB | 5010 hCoV-19/Bangladesh/DNAS_DRICM_isl_SM-12/2020\|EPI_I | ···················· | ···················· | ···················· | ···················· | ···················· | ···················· | |
| AB | 5011 hCoV-19/Bangladesh/DNAS_DRICM_isl_Sharmin/2020\|EPI | ···················· | ···················· | ···················· | ···················· | ···················· | ···················· | |
| AB | 5012 hCoV-19/Canada/ON-PHL-20-03758/2020\|EPI_ISL_775041 | ···················· | ·A·················· | ···················· | ···················· | ···················· | ··A················· | |
| AB | 5013 hCoV-19/Canada/ON-PHL-20-03782/2020\|EPI_ISL_775043 | ···················· | ·A·················· | ···················· | ···················· | ···················· | ··A················· | |
| AB | 5014 hCoV-19/Canada/ON-PHL-20-03783/2020\|EPI_ISL_775044 | ···················· | ·A·················· | ···················· | ···················· | ···················· | ··A················· | |
| AB | 5015 hCoV-19/Canada/ON-PHL-20-03784/2020\|EPI_ISL_775045 | ···················· | ·A·················· | ···················· | ···················· | ···················· | ··A················· | |
| AB | 5016 hCoV-19/Canada/ON-PHL-20-03785/2020\|EPI_ISL_775046 | ···················· | ·A·················· | ···················· | ···················· | ···················· | ··A················· | |
| AB | 5017 hCoV-19/Canada/ON-PHL-20-03786/2020\|EPI_ISL_775047 | ···················· | ·A·················· | ···················· | ···················· | ···················· | ··A················· | |
| AB | 5018 hCoV-19/Canada/ON-PHL-20-03787/2020\|EPI_ISL_775048 | ···················· | ·A·················· | ···················· | ···················· | ···················· | ··A················· | |
| AB | 5019 hCoV-19/Canada/ON-PHL-20-03788/2020\|EPI_ISL_775049 | ···················· | ·A·················· | ···················· | ···················· | ···················· | ··A················· | |
| AB | 5020 hCoV-19/Canada/ON-PHL-20-03790/2020\|EPI_ISL_775051 | ···················· | ···················· | ···················· | ···················· | ···················· | ···················· | |
| AB | 5021 hCoV-19/Canada/ON-PHL-20-03791/2020\|EPI_ISL_775052 | ···················· | ·A·················· | ···················· | ···················· | ···················· | ··A················· | |
| AB | 5022 hCoV-19/Canada/ON-PHL-20-03792/2020\|EPI_ISL_775053 | ···················· | ·A·················· | ···················· | ···················· | ···················· | ··A················· | |
| AB | 5023 hCoV-19/Canada/ON-PHL-20-03793/2020\|EPI_ISL_775054 | ···················· | ·A·················· | ···················· | ···················· | ···················· | ··A················· | |
| AB | 5024 hCoV-19/Canada/ON-PHL-20-03794/2020\|EPI_ISL_775055 | ···················· | ·A·················· | ···················· | ···················· | ···················· | ··A················· | |
| AB | 5025 hCoV-19/Canada/ON-PHL-20-03795/2020\|EPI_ISL_775056 | ···················· | ···················· | ···················· | ···················· | ···················· | ···················· | |
| AB | 5026 hCoV-19/Canada/ON-PHL-20-03796/2020\|EPI_ISL_775057 | ···················· | ···················· | ···················· | ···················· | ···················· | ···················· | |
| AB | 5027 hCoV-19/Canada/ON-PHL-20-03797/2020\|EPI_ISL_775058 | ···················· | ···················· | ···················· | ···················· | ···················· | ···················· | |
| AB | 5028 hCoV-19/Canada/ON-PHL-20-03798/2020\|EPI_ISL_775059 | ···················· | ···················· | ···················· | ···················· | ···················· | ···················· | |
| AB | 5029 hCoV-19/Canada/ON-PHL-20-03799/2020\|EPI_ISL_775060 | ···················· | ·A·················· | ···················· | ···················· | ···················· | ··A················· | |
| AB | 5030 hCoV-19/Canada/ON-PHL-20-03801/2020\|EPI_ISL_775062 | ···················· | ···················· | ···················· | ···················· | ···················· | ···················· | |
| AB | 5031 hCoV-19/Canada/ON-PHL-20-03802/2020\|EPI_ISL_775063 | ···················· | ·A·················· | ···················· | ···················· | ···················· | ··A················· | |
| AB | 5032 hCoV-19/Canada/ON-PHL-20-03803/2020\|EPI_ISL_775064 | ···················· | ·A·················· | ···················· | ···················· | ···················· | ··A················· | |
| AB | 5033 hCoV-19/Canada/ON-PHL-20-03804/2020\|EPI_ISL_775065 | ···················· | ···················· | ···················· | ···················· | ···················· | ···················· | |
| AB | 5034 hCoV-19/Canada/ON-PHL-20-03805/2020\|EPI_ISL_775066 | ···················· | ···················· | ···················· | ···················· | ···················· | ···················· | |
| AB | 5035 hCoV-19/Canada/ON-PHL-20-03806/2020\|EPI_ISL_775067 | ···················· | ···················· | ···················· | ···················· | ···················· | ···················· | |
| AB | 5036 hCoV-19/Canada/ON-PHL-20-03807/2020\|EPI_ISL_775068 | ···················· | ···················· | ···················· | ···················· | ···················· | ···················· | |
| AB | 5037 hCoV-19/Canada/ON-PHL-20-03808/2020\|EPI_ISL_775069 | ···················· | ···················· | ···················· | ···················· | ···················· | ···················· | |
| AB | 5038 hCoV-19/Canada/ON-PHL-20-03760/2020\|EPI_ISL_775070 | ···················· | ·A·················· | ···················· | ···················· | NNN················· | ··A················· | 1 |
| AB | 5039 hCoV-19/Canada/ON-PHL-20-03761/2020\|EPI_ISL_775071 | ···················· | ·A·················· | ···················· | ···················· | ···················· | ··A················· | |
| AB | 5041 hCoV-19/Canada/ON-PHL-20-03763/2020\|EPI_ISL_775073 | ···················· | ·A·················· | ···················· | ···················· | ···················· | ··A················· | |
| AB | 5042 hCoV-19/Canada/ON-PHL-20-03764/2020\|EPI_ISL_775074 | ···················· | ·A·················· | ···················· | ···················· | ···················· | ··A················· | |
| AB | 5043 hCoV-19/Canada/ON-PHL-20-03765/2020\|EPI_ISL_775075 | ···················· | ·A·················· | ···················· | ···················· | ···················· | ··A················· | |
| AB | 5044 hCoV-19/Canada/ON-PHL-20-03766/2020\|EPI_ISL_775076 | ···················· | ·A·················· | ···················· | ···················· | ···················· | ··A················· | |
| AB | 5045 hCoV-19/Canada/ON-PHL-20-03767/2020\|EPI_ISL_775077 | ···················· | ·A·················· | ···················· | ···················· | ···················· | ··A················· | |
| AB | 5046 hCoV-19/Canada/ON-PHL-20-03768/2020\|EPI_ISL_775078 | ···················· | ·A·················· | ···················· | ···················· | ···················· | ··A················· | |
| AB | 5047 hCoV-19/Canada/ON-PHL-20-03770/2020\|EPI_ISL_775079 | ···················· | ·A·················· | ···················· | ···················· | ···················· | ··A················· | |
| AB | 5048 hCoV-19/Germany/HH-hpi-p2023/2020\|EPI_ISL_776327\|2 | ···················· | ···················· | ···················· | ···················· | ···················· | ···················· | |
| AB | 5049 hCoV-19/Germany/HH-hpi-p2024/2020\|EPI_ISL_776328\|2 | ···················· | ···················· | ···················· | ···················· | ···················· | ···················· | |
| AB | 5050 hCoV-19/Germany/HH-hpi-p2027/2020\|EPI_ISL_776329\|2 | ···················· | ···················· | ···················· | ···················· | ···················· | ···················· | |
| AB | 5051 hCoV-19/Germany/HH-hpi-p2048/2020\|EPI_ISL_776338\|2 | ···················· | ···················· | ···················· | ···················· | ···················· | ···················· | |
| AB | 5052 hCoV-19/Germany/HH-hpi-p2049/2020\|EPI_ISL_776339\|2 | ···················· | ···················· | ···················· | ···················· | ···················· | ···················· | |
| AB | 5053 hCoV-19/Germany/HH-hpi-p2051/2020\|EPI_ISL_776340\|2 | ···················· | ···················· | ···················· | ···················· | ···················· | ···················· | |
| AB | 5054 hCoV-19/Germany/HH-hpi-p2053/2020\|EPI_ISL_776341\|2 | ···················· | ···················· | ···················· | ···················· | ···················· | ···················· | |
| AB | 5055 hCoV-19/Germany/HH-hpi-p2054/2020\|EPI_ISL_776342\|2 | ···················· | ···················· | ···················· | ···················· | ···················· | ···················· | |
| AB | 5056 hCoV-19/Germany/HH-hpi-p2056/2020\|EPI_ISL_776343\|2 | ···················· | ···················· | ···················· | ···················· | ···················· | ···················· | |
| AB | 5057 hCoV-19/Germany/HH-hpi-p2060/2020\|EPI_ISL_776344\|2 | ···················· | ···················· | ···················· | ···················· | ···················· | ···················· | |
| AB | 5058 hCoV-19/Germany/HH-hpi-p2061/2020\|EPI_ISL_776345\|2 | ···················· | ···················· | ···················· | ···················· | ···················· | ···················· | |
| AB | 5059 hCoV-19/Germany/HH-hpi-p2063/2020\|EPI_ISL_776346\|2 | ···················· | ···················· | ···················· | ···················· | ···················· | ···················· | |
| AB | 5060 hCoV-19/Germany/HH-hpi-p2071/2020\|EPI_ISL_776347\|2 | ···················· | ···················· | ···················· | ···················· | ···················· | ···················· | |
| AB | 5061 hCoV-19/Germany/HH-hpi-p2076/2020\|EPI_ISL_776348\|2 | ···················· | ···················· | ···················· | ···················· | ···················· | ···················· | |
| AB | 5062 hCoV-19/Germany/HH-hpi-p2130/2020\|EPI_ISL_776370\|2 | ···················· | ···················· | ···················· | ···················· | ···················· | ···················· | |
| AB | 5063 hCoV-19/Germany/HH-hpi-p2131/2020\|EPI_ISL_776371\|2 | ···················· | ···················· | ···················· | ···················· | ···················· | ···················· | |
| AB | 5064 hCoV-19/Germany/HH-hpi-p2133/2020\|EPI_ISL_776372\|2 | ···················· | ···················· | ···················· | ···················· | ···················· | ···················· | |
| AB | 5065 hCoV-19/Germany/HH-hpi-p2135/2020\|EPI_ISL_776373\|2 | ···················· | ···················· | ···················· | ···················· | ···················· | ···················· | |
| AB | 5066 hCoV-19/Germany/HH-hpi-p2136/2020\|EPI_ISL_776374\|2 | ···················· | ···················· | ···················· | ···················· | ···················· | ···················· | |
| AB | 5067 hCoV-19/Germany/HH-hpi-p2138/2020\|EPI_ISL_776375\|2 | ···················· | ···················· | ···················· | ···················· | ···················· | ···················· | |
| AB | 5068 hCoV-19/Germany/HH-hpi-p2144/2020\|EPI_ISL_776376\|2 | ···················· | ···················· | ···················· | ···················· | ···················· | ···················· | |

| | | | | | | | | |
|---|---|---|---|---|---|---|---|---|
| AB | 5069 | hCoV-19/Germany/HH-hpi-p2145/2020\|EPI_ISL_776377\|2 | ···················· | ····A··············· | ···················· | ···················· | ···················· | ···················· |
| AB | 5070 | hCoV-19/Germany/HH-hpi-p2146/2020\|EPI_ISL_776378\|2 | ···················· | ···················· | ···················· | ···················· | ···················· | ···················· |
| AB | 5071 | hCoV-19/Germany/HH-hpi-p2147/2020\|EPI_ISL_776379\|2 | ···················· | ···················· | ···················· | ···················· | ···················· | ···················· |
| AB | 5072 | hCoV-19/Germany/HH-hpi-p2148/2020\|EPI_ISL_776380\|2 | ···················· | ···················· | ···················· | ···················· | ···················· | ···················· |
| AB | 5073 | hCoV-19/Germany/HH-hpi-p2149/2020\|EPI_ISL_776381\|2 | ···················· | ···················· | ···················· | ···················· | ···················· | ···················· |
| AB | 5074 | hCoV-19/Germany/HH-hpi-p2150/2020\|EPI_ISL_776382\|2 | ···················· | ···················· | ···················· | ···················· | ···················· | ···················· |
| AB | 5075 | hCoV-19/Germany/HH-hpi-p2151/2020\|EPI_ISL_776383\|2 | ···················· | ···················· | ···················· | ···················· | ···················· | ···················· |
| AB | 5076 | hCoV-19/Germany/HH-hpi-p2152/2020\|EPI_ISL_776384\|2 | ···················· | ···················· | ···················· | ···················· | ···················· | ···················· |
| AB | 5077 | hCoV-19/Germany/HH-hpi-p2153/2020\|EPI_ISL_776385\|2 | ···················· | ···················· | ···················· | ···················· | ···················· | ···················· |
| AB | 5078 | hCoV-19/Germany/HH-hpi-p2154/2020\|EPI_ISL_776386\|2 | ···················· | ···················· | ···················· | ···················· | ···················· | ···················· |
| AB | 5079 | hCoV-19/Germany/HH-hpi-p2155/2020\|EPI_ISL_776387\|2 | ···················· | ···················· | ···················· | ···················· | ···················· | ···················· |
| AB | 5080 | hCoV-19/Germany/HH-hpi-p2156/2020\|EPI_ISL_776388\|2 | ···················· | ···················· | ···················· | ···················· | ···················· | ···················· |
| AB | 5081 | hCoV-19/Germany/HH-hpi-p2157/2020\|EPI_ISL_776389\|2 | ···················· | ···················· | ···················· | ···················· | ···················· | ···················· |
| AB | 5082 | hCoV-19/Germany/HH-hpi-p2158/2020\|EPI_ISL_776390\|2 | ···················· | ···················· | ···················· | ···················· | ···················· | ···················· |
| AB | 5083 | hCoV-19/Germany/HH-hpi-p2159/2020\|EPI_ISL_776391\|2 | ···················· | ···················· | ···················· | ···················· | ···················· | ···················· |
| AB | 5084 | hCoV-19/Germany/HH-hpi-p2160/2020\|EPI_ISL_776392\|2 | ···················· | ···················· | ···················· | ···················· | ···················· | ···················· |
| AB | 5085 | hCoV-19/Germany/HH-hpi-p2163/2020\|EPI_ISL_776393\|2 | ···················· | ···················· | ···················· | ···················· | ···················· | ···················· |
| AB | 5086 | hCoV-19/Germany/HH-hpi-p2164/2020\|EPI_ISL_776394\|2 | ···················· | ···················· | ···················· | ···················· | ···················· | ···················· |
| AB | 5087 | hCoV-19/Germany/HH-hpi-p2165/2020\|EPI_ISL_776395\|2 | ···················· | ···················· | ···················· | ···················· | ···················· | ···················· |
| AB | 5088 | hCoV-19/Germany/HH-hpi-p2166/2020\|EPI_ISL_776396\|2 | ···················· | ···················· | ···················· | ···················· | ···················· | ···················· |
| AB | 5089 | hCoV-19/Germany/HH-hpi-p2167/2020\|EPI_ISL_776397\|2 | ···················· | ···················· | ···················· | ···················· | ···················· | ···················· |
| AB | 5090 | hCoV-19/Germany/HH-hpi-p2168/2020\|EPI_ISL_776398\|2 | ···················· | ···················· | ···················· | ···················· | ···················· | ···················· |
| AB | 5091 | hCoV-19/Germany/HH-hpi-p2169/2020\|EPI_ISL_776399\|2 | ···················· | ···················· | ···················· | ···················· | ···················· | ···················· |
| AB | 5092 | hCoV-19/England/QEUH-C93E66/2020\|EPI_ISL_777600\|20 | ···················· | ···················· | ···················· | ···················· | ···················· | ···················· |
| AB | 5093 | hCoV-19/England/QEUH-C93E39/2020\|EPI_ISL_777601\|20 | ···················· | ···················· | ···················· | ···················· | ···················· | ···················· |
| AB | 5094 | hCoV-19/England/CAMC-C9849C/2020\|EPI_ISL_777609\|20 | ···················· | ···················· | ···················· | ···················· | ···················· | ···················· |
| AB | 5095 | hCoV-19/England/ALDP-C8E6C0/2020\|EPI_ISL_777672\|20 | ···················· | ···················· | ···················· | ···················· | ···················· | ···················· |
| AB | 5096 | hCoV-19/England/CAMC-CB79D4/2020\|EPI_ISL_777679\|20 | ···················· | ···················· | ···················· | ···················· | ···················· | ···················· |
| AB | 5097 | hCoV-19/USA/MD-MDH-0535/2020\|EPI_ISL_778911\|2020-1 | ···················· | ···················· | ···················· | ···················· | ···················· | ···················· |
| AB | 5098 | hCoV-19/USA/LA-EVTL1257/2020\|EPI_ISL_778923\|2020-1 | ···················· | ···················· | ···················· | ···················· | ···················· | ···················· |
| AB | 5099 | hCoV-19/USA/LA-EVTL1258/2020\|EPI_ISL_778924\|2020-1 | ···················· | ···················· | ···················· | ···················· | ···················· | ···················· |
| AB | 5100 | hCoV-19/USA/LA-EVTL1259/2020\|EPI_ISL_778925\|2020-1 | ···················· | ···················· | ···················· | ···················· | ···················· | ···················· |
| AB | 5101 | hCoV-19/USA/LA-EVTL1260/2020\|EPI_ISL_778926\|2020-1 | ···················· | ···················· | ···················· | ···················· | ···················· | ···················· |
| AB | 5102 | hCoV-19/USA/LA-EVTL1261/2020\|EPI_ISL_778927\|2020-1 | ···················· | ···················· | ···················· | ···················· | ···················· | ···················· |
| AB | 5103 | hCoV-19/USA/LA-EVTL1262/2020\|EPI_ISL_778928\|2020-1 | ···················· | ···················· | ···················· | ···················· | ···················· | ···················· |
| AB | 5104 | hCoV-19/USA/LA-EVTL1263/2020\|EPI_ISL_778929\|2020-1 | ···················· | ···················· | ···················· | ···················· | ···················· | ···················· |
| AB | 5105 | hCoV-19/USA/LA-EVTL1264/2020\|EPI_ISL_778930\|2020-1 | ···················· | ···················· | ···················· | ···················· | ···················· | ···················· |
| AB | 5106 | hCoV-19/USA/LA-EVTL1265/2020\|EPI_ISL_778931\|2020-1 | ···················· | ···················· | ···················· | ···················· | ···················· | ···················· |
| AB | 5107 | hCoV-19/USA/LA-EVTL1266/2020\|EPI_ISL_778932\|2020-1 | ···················· | ···················· | ···················· | ···················· | ···················· | ···················· |
| AB | 5108 | hCoV-19/USA/LA-EVTL1267/2020\|EPI_ISL_778933\|2020-1 | ···················· | ···················· | ···················· | ···················· | ···················· | ···················· |
| AB | 5109 | hCoV-19/USA/LA-EVTL1268/2020\|EPI_ISL_778934\|2020-1 | ···················· | ···················· | ···················· | ···················· | ···················· | ···················· |
| AB | 5110 | hCoV-19/USA/LA-EVTL1269/2020\|EPI_ISL_778935\|2020-1 | ···················· | ···················· | ···················· | ···················· | ···················· | ···················· |
| AB | 5111 | hCoV-19/USA/LA-EVTL1270/2020\|EPI_ISL_778936\|2020-1 | ···················· | ···················· | ···················· | ···················· | ···················· | ···················· |
| AB | 5112 | hCoV-19/USA/LA-EVTL1271/2020\|EPI_ISL_778937\|2020-1 | ···················· | ···················· | ···················· | ···················· | ···················· | ···················· |
| AB | 5113 | hCoV-19/USA/LA-EVTL1272/2020\|EPI_ISL_778938\|2020-1 | ···················· | ···················· | ···················· | ···················· | ···················· | ···················· |
| AB | 5114 | hCoV-19/USA/LA-EVTL1273/2020\|EPI_ISL_778939\|2020-1 | ···················· | ···················· | ···················· | ···················· | ···················· | ···················· |
| AB | 5115 | hCoV-19/USA/LA-EVTL1274/2020\|EPI_ISL_778940\|2020-1 | ···················· | ···················· | ···················· | ···················· | ···················· | ···················· |
| AB | 5116 | hCoV-19/USA/LA-EVTL1275/2020\|EPI_ISL_778941\|2020-1 | ···················· | ···················· | ···················· | ···················· | ···················· | ···················· |
| AB | 5117 | hCoV-19/USA/LA-EVTL1276/2020\|EPI_ISL_778942\|2020-1 | ···················· | ···················· | ···················· | ···················· | ···················· | ···················· |
| AB | 5118 | hCoV-19/USA/LA-EVTL1277/2020\|EPI_ISL_778943\|2020-1 | ···················· | ···················· | ···················· | ···················· | ···················· | ···················· |
| AB | 5119 | hCoV-19/USA/LA-EVTL1278/2020\|EPI_ISL_778944\|2020-1 | ···················· | ···················· | ···················· | ···················· | ···················· | ···················· |
| AB | 5120 | hCoV-19/USA/LA-EVTL1323/2020\|EPI_ISL_778989\|2020-1 | ···················· | ···················· | ···················· | ···················· | ···················· | ···················· |
| AB | 5121 | hCoV-19/Finland/24SS6bF7/2020\|EPI_ISL_759838\|2020- | ···················· | ·A·················· | ···················· | ···················· | ···················· | ···················· |
| AB | 5122 | hCoV-19/Finland/24SS6E1/2020\|EPI_ISL_759839\|2020-0 | ···················· | ···················· | ···················· | ···················· | ···················· | ···················· |
| AB | 5123 | hCoV-19/Finland/24SS7B9/2020\|EPI_ISL_759840\|2020-0 | ···················· | ·A·················· | ···················· | ···················· | ···················· | ···A················ |
| AB | 5124 | hCoV-19/Finland/24SS9A5/2020\|EPI_ISL_759841\|2020-0 | ···················· | ·A·················· | ···················· | ···················· | ···················· | ···A················ |
| AB | 5125 | hCoV-19/Vietnam/40219/2020\|EPI_ISL_759869\|2020-08 | ···················· | ···················· | ···················· | ···················· | ···················· | ···················· |
| AB | 5126 | hCoV-19/Vietnam/40228/2020\|EPI_ISL_759870\|2020-08 | ···················· | ···················· | ···················· | ···················· | ···················· | ···················· |
| AB | 5127 | hCoV-19/Vietnam/40227/2020\|EPI_ISL_759871\|2020-08 | ···················· | ···················· | ···················· | ···················· | ···················· | ···················· |
| AB | 5128 | hCoV-19/Vietnam/40229/2020\|EPI_ISL_759872\|2020-08 | ···················· | ···················· | ···················· | ···················· | ···················· | ···················· |
| AB | 5129 | hCoV-19/Vietnam/40113/2020\|EPI_ISL_759873\|2020-07 | ···················· | ···················· | ···················· | ···················· | ···················· | ···················· |
| AB | 5130 | hCoV-19/Vietnam/40124/2020\|EPI_ISL_759874\|2020-07 | ···················· | ···················· | ···················· | ···················· | ···················· | ···················· |
| AB | 5131 | hCoV-19/Vietnam/40125/2020\|EPI_ISL_759875\|2020-07 | ···················· | ···················· | ···················· | ···················· | ···················· | ···················· |
| AB | 5132 | hCoV-19/Vietnam/40126/2020\|EPI_ISL_759876\|2020-07 | ···················· | ···················· | ···················· | ···················· | ···················· | ···················· |
| AB | 5133 | hCoV-19/Vietnam/40127/2020\|EPI_ISL_759877\|2020-07 | ···················· | ···················· | ···················· | ···················· | ···················· | ···················· |
| AB | 5134 | hCoV-19/Vietnam/40128/2020\|EPI_ISL_759878\|2020-07 | ···················· | ···················· | ···················· | ···················· | ···················· | ···················· |
| AB | 5135 | hCoV-19/Vietnam/40130/2020\|EPI_ISL_759879\|2020-07 | ···················· | ···················· | ···················· | ···················· | ···················· | ···················· |
| AB | 5136 | hCoV-19/Vietnam/40131/2020\|EPI_ISL_759880\|2020-07 | ···················· | ···················· | ···················· | ···················· | ···················· | ···················· |
| AB | 5137 | hCoV-19/Vietnam/40188/2020\|EPI_ISL_759881\|2020-07 | ···················· | ···················· | ···················· | ···················· | ···················· | ···················· |
| AB | 5138 | hCoV-19/Vietnam/40191/2020\|EPI_ISL_759882\|2020-08 | ···················· | ···················· | ···················· | ···················· | ···················· | ···················· |
| AB | 5139 | hCoV-19/Vietnam/40201/2020\|EPI_ISL_759883\|2020-08 | ···················· | ···················· | ···················· | ···················· | ···················· | ···················· |
| AB | 5140 | hCoV-19/Vietnam/40203/2020\|EPI_ISL_759884\|2020-08 | ···················· | ···················· | ···················· | ···················· | ···················· | ···················· |
| AB | 5141 | hCoV-19/Vietnam/40205/2020\|EPI_ISL_759885\|2020-08 | ···················· | ···················· | ···················· | ···················· | ···················· | ···················· |
| AB | 5142 | hCoV-19/Vietnam/40213/2020\|EPI_ISL_759886\|2020-07 | ···················· | ···················· | ···················· | ···················· | ···················· | ···················· |
| AB | 5143 | hCoV-19/Vietnam/40218/2020\|EPI_ISL_759887\|2020-08 | ···················· | ···················· | ···················· | ···················· | ···················· | ···················· |
| AB | 5144 | hCoV-19/Vietnam/40230/2020\|EPI_ISL_759888\|2020-08 | ···················· | ···················· | ···················· | ···················· | ···················· | ···················· |
| AB | 5145 | hCoV-19/Vietnam/40234/2020\|EPI_ISL_759889\|2020-08 | ···················· | ···················· | ···················· | ···················· | ···················· | ···················· |
| AB | 5146 | hCoV-19/Vietnam/40236/2020\|EPI_ISL_759890\|2020-08 | ···················· | ···················· | ···················· | ···················· | ···················· | ···················· |
| AB | 5147 | hCoV-19/Vietnam/CM18609/2020\|EPI_ISL_759891\|2020-0 | ···················· | ···················· | ···················· | ···················· | ···················· | ···················· |
| AB | 5148 | hCoV-19/HongKong/CM20000489/2020\|EPI_ISL_759892\|20 | ···················· | ···················· | ···················· | ···················· | ···················· | ···················· |

AB 5149 hCoV-19/HongKong/CM20000494/2020|EPI_ISL_759893|20 ···················· ···················· ···················· ···················· ···················· ····················
AB 5150 hCoV-19/HongKong/CM20000499/2020|EPI_ISL_759894|20 ···················· ···················· ···················· ···················· ···················· ····················
AB 5151 hCoV-19/HongKong/VM20194998/2020|EPI_ISL_759895|20 ···················· ·A·················· ···················· ···················· ···················· ··A·················
AB 5152 hCoV-19/HongKong/VM20194997/2020|EPI_ISL_759897|20 ···················· ·A·················· ···················· ···················· ···················· ··A·················
AB 5153 hCoV-19/HongKong/VM20194999/2020|EPI_ISL_759898|20 ···················· ·A·················· ···················· ···················· ···················· ··A·················
AB 5154 hCoV-19/Finland/25ES3AB11/2020|EPI_ISL_759899|2020 ···················· ·A·················· ···················· ···················· ···················· ··A·················
AB 5155 hCoV-19/England/QEUH-CADD6C/2020|EPI_ISL_760820|20 ···················· ···················· ···················· ···················· ·····T·············· ····················
AB 5156 hCoV-19/England/CAMC-C1CFDB/2020|EPI_ISL_760822|20 ···················· ···················· ···················· ···················· ···················· ····················
AB 5157 hCoV-19/England/QEUH-CADE5A/2020|EPI_ISL_760823|20 ···················· ···················· ···················· ···················· ···················· T···················
AB 5158 hCoV-19/England/QEUH-CAD24F/2020|EPI_ISL_760824|20 ···················· ···················· ···················· ···················· ···················· ····················
AB 5159 hCoV-19/England/ALDP-CB3D54/2020|EPI_ISL_760826|20 ···················· ···················· ···················· ···················· ···················· ····················
AB 5160 hCoV-19/England/QEUH-CAD898/2020|EPI_ISL_760828|20 ···················· ···················· ···················· ···················· ···················· ····················
AB 5161 hCoV-19/England/ALDP-CB3D81/2020|EPI_ISL_760832|20 ···················· ···················· ···················· ···················· ···················· ··A·················
AB 5162 hCoV-19/England/CAMC-CB2D73/2020|EPI_ISL_760833|20 ···················· ·A·················· ···················· ···················· ···················· ··A·················
AB 5163 hCoV-19/England/ALDP-CB3D18/2020|EPI_ISL_760838|20 ···················· ···················· ···················· ···················· ···················· ····················
AB 5165 hCoV-19/England/QEUH-CAB618/2020|EPI_ISL_760840|20 ···················· ···················· ···················· ···················· ···················· ····················
AB 5166 hCoV-19/England/QEUH-CAD8B6/2020|EPI_ISL_760841|20 ···················· ···················· ···················· ···················· ···················· ····················
AB 5167 hCoV-19/England/QEUH-CAA52B/2020|EPI_ISL_760843|20 ···················· ···················· ···················· ···················· ···················· ····················
AB 5168 hCoV-19/England/MILK-CB1CA4/2020|EPI_ISL_760844|20 ···················· ·A·················· ···················· ···················· ···················· ····················
AB 5169 hCoV-19/England/QEUH-CADEB4/2020|EPI_ISL_760845|20 ···················· ···················· ···················· ···················· ·····T·············· ····················
AB 5170 hCoV-19/England/QEUH-CAEA82/2020|EPI_ISL_760847|20 ···················· ···················· ···················· ···················· ···················· ····················
AB 5171 hCoV-19/England/QEUH-CAD3B5/2020|EPI_ISL_760848|20 ···················· ···················· ···················· ···················· ···················· ····················
AB 5172 hCoV-19/England/QEUH-CABE01/2020|EPI_ISL_760849|20 ···················· ···················· ···················· ···················· ···················· ····················
AB 5173 hCoV-19/England/QEUH-CADA74/2020|EPI_ISL_760851|20 ···················· ···················· ···················· ···················· ···················· ····················
AB 5174 hCoV-19/England/QEUH-CAA5D0/2020|EPI_ISL_760854|20 ···················· ·A·················· ···················· ···················· ···················· ··A·················
AB 5175 hCoV-19/England/QEUH-CAD0BE/2020|EPI_ISL_760855|20 ···················· ·A·················· ···················· ···················· ···················· ··A·················
AB 5176 hCoV-19/England/QEUH-CAD519/2020|EPI_ISL_760856|20 ···················· ···················· ···················· ···················· ···················· ····················
AB 5177 hCoV-19/England/ALDP-CB4755/2020|EPI_ISL_760857|20 ···················· ···················· ···················· ···················· ···················· ····················
AB 5178 hCoV-19/England/CAMC-CB2BD3/2020|EPI_ISL_760858|20 ···················· ···················· ···················· ···················· ···················· ····················
AB 5179 hCoV-19/England/CAMC-CAF975/2020|EPI_ISL_760859|20 ···················· ···················· ···················· ···················· ···················· ····················
AB 5180 hCoV-19/England/ALDP-CB46D0/2020|EPI_ISL_760861|20 ···················· ···················· ···················· ···················· ·····T·············· ····················
AB 5181 hCoV-19/England/ALDP-CB3F5E/2020|EPI_ISL_760862|20 ···················· ···················· ···················· ···················· ···················· ····················
AB 5182 hCoV-19/England/MILK-CB1C86/2020|EPI_ISL_760863|20 ···················· ···················· ···················· ···················· ···················· ····················
AB 5183 hCoV-19/England/ALDP-CB47CE/2020|EPI_ISL_760864|20 ···················· ···················· ···················· ···················· ···················· ····················
AB 5184 hCoV-19/England/ALDP-CB45C4/2020|EPI_ISL_760865|20 ···················· ···················· ···················· ···················· ·····T·············· ····················
AB 5185 hCoV-19/England/QEUH-C02891/2020|EPI_ISL_760866|20 ···················· ···················· ···················· ···················· ···················· ····················
AB 5186 hCoV-19/England/ALDP-CB4227/2020|EPI_ISL_760867|20 ···················· ···················· ···················· ···················· ···················· ····················
AB 5187 hCoV-19/England/ALDP-CB4773/2020|EPI_ISL_760868|20 ···················· ···················· ···················· ···················· ···················· ····················
AB 5188 hCoV-19/England/QEUH-CB0D66/2020|EPI_ISL_760872|20 ···················· ·A·················· ···················· ···················· ···················· ··A·················
AB 5189 hCoV-19/England/MILK-CB067A/2020|EPI_ISL_760873|20 ···················· ···················· ···················· ···················· ···················· ····················
AB 5190 hCoV-19/England/QEUH-CADAA1/2020|EPI_ISL_760875|20 ···················· ···················· ···················· ···················· ···················· ····················
AB 5191 hCoV-19/England/QEUH-CAD0AF/2020|EPI_ISL_760879|20 ···················· ···················· ···················· ···················· ·····T·············· ····················
AB 5192 hCoV-19/England/QEUH-CADA92/2020|EPI_ISL_760882|20 ···················· ···················· ···················· ···················· ···················· ····················
AB 5193 hCoV-19/England/ALDP-CB4245/2020|EPI_ISL_760884|20 ···················· ···················· ···················· ···················· ···················· ····················
AB 5194 hCoV-19/England/QEUH-CAD9B3/2020|EPI_ISL_760885|20 ···················· ···················· ···················· ···················· ···················· ····················
AB 5195 hCoV-19/England/ALDP-CB470A/2020|EPI_ISL_760887|20 ···················· ···················· ···················· ···················· ···················· ····················
AB 5196 hCoV-19/England/ALDP-CB3D09/2020|EPI_ISL_760890|20 ···················· ···················· ···················· ···················· ···················· ····················
AB 5197 hCoV-19/England/ALDP-CB30B5/2020|EPI_ISL_760891|20 ···················· ···················· ···················· ···················· ···················· ····················
AB 5198 hCoV-19/England/ALDP-CB462B/2020|EPI_ISL_760892|20 ···················· ···················· ···················· ···················· ·····T·············· ····················
AB 5199 hCoV-19/England/CAMC-C1098E/2020|EPI_ISL_760894|20 ···················· ···················· ···················· ···················· ···················· ····················
AB 5200 hCoV-19/England/MILK-CB1BC5/2020|EPI_ISL_760896|20 ···················· ···················· ···················· ···················· ···················· ····················
AB 5201 hCoV-19/England/ALDP-CB4597/2020|EPI_ISL_760897|20 ···················· ···················· ···················· ···················· ···················· ····················
AB 5202 hCoV-19/Germany/NW-RKI-NP-0005/2020|EPI_ISL_763028 ···················· ···················· ···················· ···················· ···················· ····················
AB 5204 hCoV-19/Germany/HE-RKI-NP-0053/2020|EPI_ISL_763031 ···················· ···················· ···················· ···················· ···················· ····················
AB 5205 hCoV-19/Germany/HE-RKI-NP-0008/2020|EPI_ISL_763034 ···················· ···················· ···················· ···················· ···················· ····················
AB 5206 hCoV-19/Germany/BY-RKI-NP-0059/2020|EPI_ISL_763037 ···················· ···················· ···················· ···················· ···················· ····················
AB 5208 hCoV-19/Germany/BW-RKI-NP-0052/2020|EPI_ISL_763040 ···················· ···················· ···················· ···················· ···················· ····················
AB 5209 hCoV-19/Germany/BW-RKI-NP-0051/2020|EPI_ISL_763041 ···················· ···················· ···················· ···················· ···················· ····················
AB 5210 hCoV-19/Germany/BW-RKI-NP-0040/2020|EPI_ISL_763043 ···················· ···················· ···················· ···················· ···················· ····················
AB 5212 hCoV-19/Germany/BW-RKI-NP-0025/2020|EPI_ISL_763045 ···················· ···················· ···················· ···················· ···················· ····················
AB 5213 hCoV-19/Germany/BW-RKI-NP-0023/2020|EPI_ISL_763046 ···················· ···················· ···················· ···················· ···················· ····················
AB 5214 hCoV-19/Germany/BW-RKI-NP-0020/2020|EPI_ISL_763047 ···················· ···················· ···················· ···················· ···················· ····················
AB 5215 hCoV-19/Germany/BW-RKI-NP-0016/2020|EPI_ISL_763048 ···················· ···················· ···················· ···················· ···················· ····················
AB 5217 hCoV-19/Germany/BW-RKI-NP-0009/2020|EPI_ISL_763050 ···················· ···················· ···················· ···················· ···················· ····················
AB 5218 hCoV-19/Germany/BW-RKI-NP-0002/2020|EPI_ISL_763051 ···················· ···················· ···················· ···················· ···················· ····················
AB 5219 hCoV-19/Germany/BB-RKI-NP-0068/2020|EPI_ISL_763053 ···················· ···················· ···················· ···················· ···················· ····················
AB 5220 hCoV-19/Germany/BB-RKI-NP-0067/2020|EPI_ISL_763054 ···················· ···················· ···················· ···················· ···················· ····················
AB 5221 hCoV-19/Germany/BB-RKI-NP-0058/2020|EPI_ISL_763055 ···················· ···················· ···················· ···················· ···················· ····················
AB 5222 hCoV-19/Germany/BB-RKI-NP-0057/2020|EPI_ISL_763056 ···················· ···················· ···················· ···················· ···················· ····················
AB 5223 hCoV-19/Germany/BB-RKI-NP-0056/2020|EPI_ISL_763057 ···················· ···················· ···················· ···················· ···················· ····················
AB 5224 hCoV-19/Germany/BB-RKI-NP-0055/2020|EPI_ISL_763058 ···················· ···················· ···················· ···················· ···················· ····················
AB 5225 hCoV-19/Germany/BB-RKI-NP-0038/2020|EPI_ISL_763059 ···················· ···················· ···················· ···················· ···················· ····················
AB 5226 hCoV-19/Scotland/EDB11089/2020|EPI_ISL_764359|2020 ···················· ···················· ···················· ···················· ···················· ····················
AB 5227 hCoV-19/Scotland/EDB11315/2020|EPI_ISL_764362|2020 ···················· ···················· ···················· ···················· ···················· ····················
AB 5228 hCoV-19/Scotland/EDB11317/2020|EPI_ISL_764363|2020 ···················· ···················· ···················· ···················· ···················· ····················
AB 5229 hCoV-19/Scotland/EDB11319/2020|EPI_ISL_764364|2020 ···················· ···················· ···················· ···················· ···················· ····················
AB 5230 hCoV-19/Scotland/EDB11322/2020|EPI_ISL_764365|2020 ···················· ···················· ···················· ···················· ···················· ····················
AB 5231 hCoV-19/Scotland/EDB11328/2020|EPI_ISL_764366|2020 ···················· ···················· ···················· ···················· ···················· ····················
AB 5233 hCoV-19/Scotland/EDB11336/2020|EPI_ISL_764369|2020 ···················· ···················· ···················· ···················· ···················· ····················
AB 5235 hCoV-19/Scotland/EDB11340/2020|EPI_ISL_764371|2020 ···················· ···················· ···················· ···················· ···················· ····················

```
AB  5236 hCoV-19/Scotland/EDB11341/2020|EPI_ISL_764372|2020  ···················· ···················· ···················· ···················· ···················· ····················
AB  5237 hCoV-19/England/EXET-13EBBC/2020|EPI_ISL_764373|20  ···················· ···················· ···················· ···················· ···················· ····················
AB  5238 hCoV-19/England/EXET-13ECB9/2020|EPI_ISL_764374|20  ···················· ···················· ···················· ···················· ······T············· ····················
AB  5239 hCoV-19/England/EXET-13ED7A/2020|EPI_ISL_764375|20  ···················· ···················· ···················· ···················· ···················· ····················
AB  5240 hCoV-19/England/EXET-13EE2C/2020|EPI_ISL_764376|20  ···················· ···················· ···················· ···················· ···················· ····················
AB  5241 hCoV-19/England/EXET-13EEC2/2020|EPI_ISL_764377|20  ···················· ·A·················· ···················· ···················· ······T············· ····················
AB  5242 hCoV-19/England/EXET-13F24D/2020|EPI_ISL_764378|20  ···················· ···················· ···················· ···················· ···················· ····················
AB  5243 hCoV-19/England/EXET-13F2C5/2020|EPI_ISL_764379|20  ···················· ···················· ···················· ···················· ···················· ····················
AB  5244 hCoV-19/England/EXET-13F2D4/2020|EPI_ISL_764380|20  ···················· ···················· ···················· ···················· ···················· ····················
AB  5245 hCoV-19/England/EXET-13F3D1/2020|EPI_ISL_764381|20  ···················· ···················· ···················· ···················· ···················· ····················
AB  5246 hCoV-19/England/EXET-13F429/2020|EPI_ISL_764382|20  ···················· ···················· ···················· ···················· ···················· ····················
AB  5247 hCoV-19/England/LOND-12F16B0/2020|EPI_ISL_764383|2  ···················· ···················· ···················· ···················· ···················· ····················
AB  5248 hCoV-19/England/LOND-12F16CF/2020|EPI_ISL_764384|2  ···················· ···················· ···················· ···················· ···················· ····················
AB  5249 hCoV-19/England/LOND-12F1726/2020|EPI_ISL_764385|2  ···················· ···················· ···················· ···················· ···················· ····················
AB  5250 hCoV-19/England/LOND-12F1AF0/2020|EPI_ISL_764386|2  ···················· ···················· ···················· ···················· ···················· ····················
AB  5251 hCoV-19/Northern Ireland/NIRE-234AC7/2020|EPI_ISL_  ···················· ···················· ···················· ···················· ···················· ····················
AB  5252 hCoV-19/Northern Ireland/NIRE-234AD6/2020|EPI_ISL_  ···················· ···················· ···················· ···················· ···················· ····················
AB  5253 hCoV-19/Northern Ireland/NIRE-234B2E/2020|EPI_ISL_  ···················· ···················· ···················· ···················· ···················· ····················
AB  5254 hCoV-19/USA/MA-MGH-03048/2020|EPI_ISL_765693|2020-  ···················· ···················· ···················· ···················· ···················· ····················
AB  5255 hCoV-19/USA/MA-MGH-03052/2020|EPI_ISL_765694|2020-  ···················· ···················· ···················· ···················· ···················· ····················
AB  5256 hCoV-19/USA/MA-MGH-03055/2020|EPI_ISL_765695|2020-  ···················· ···················· ···················· ···················· ···················· ····················
AB  5257 hCoV-19/USA/MA-MGH-03057/2020|EPI_ISL_765696|2020-  ···················· ···················· ···················· ···················· ···················· ····················
AB  5258 hCoV-19/USA/MA-MGH-03059/2020|EPI_ISL_765697|2020-  ···················· ···················· ···················· ·············A······ ···················· ····················
AB  5259 hCoV-19/USA/MA-MGH-03060/2020|EPI_ISL_765698|2020-  ···················· ···················· ···················· ···················· ···················· ····················
AB  5260 hCoV-19/USA/MA-MGH-03077/2020|EPI_ISL_765700|2020-  ···················· ···················· ···················· ···················· ···················· ····················
AB  5261 hCoV-19/USA/MA-MGH-03113/2020|EPI_ISL_765707|2020-  ···················· ···················· ···················· ···················· ···················· ····················
AB  5262 hCoV-19/USA/MA-MGH-03120/2020|EPI_ISL_765710|2020-  ···················· ···················· ···················· ···················· ···················· ····················
AB  5263 hCoV-19/USA/MA-MGH-03124/2020|EPI_ISL_765711|2020-  ···················· ···················· ···················· ···················· ···················· ····················
AB  5264 hCoV-19/England/CAMC-C75529/2020|EPI_ISL_769201|20  ···················· ···················· ···················· ···················· ···················· ····················
AB  5265 hCoV-19/England/CAMC-C70B68/2020|EPI_ISL_769207|20  ···················· ···················· ···················· ···················· ···················· ····················
AB  5266 hCoV-19/England/CAMC-C738E6/2020|EPI_ISL_769210|20  ···················· ···················· ···················· ···················· ······T············· ····················
AB  5267 hCoV-19/England/CAMC-C71C73/2020|EPI_ISL_769211|20  ···················· ···················· ···················· ···················· ···················· ····················
AB  5268 hCoV-19/England/CAMC-C7389B/2020|EPI_ISL_769212|20  ···················· ···················· ···················· ···················· ···················· ····················
AB  5269 hCoV-19/England/CAMC-C70CED/2020|EPI_ISL_769213|20  ···················· ···················· ···················· ···················· ···················· ····················
AB  5270 hCoV-19/England/CAMC-C7210A/2020|EPI_ISL_769214|20  ···················· ·A·················· ···················· ···················· ···················· ····················
AB  5271 hCoV-19/England/CAMC-C70C92/2020|EPI_ISL_769219|20  ···················· ···················· ···················· ···················· ···················· ····················
AB  5272 hCoV-19/England/CAMC-C74997/2020|EPI_ISL_769221|20  ···················· ···················· ···················· ···················· ···················· ····················
AB  5273 hCoV-19/England/QEUH-C5CA92/2020|EPI_ISL_769229|20  ···················· ···················· ···················· ···················· ···················· ····················
AB  5274 hCoV-19/England/MILK-C61432/2020|EPI_ISL_769230|20  ···················· ···················· ···················· ···················· ···················· ····················
AB  5275 hCoV-19/England/QEUH-C5B91E/2020|EPI_ISL_769233|20  ···················· ···················· ···················· ···················· ···················· ····················
AB  5276 hCoV-19/England/QEUH-C5C054/2020|EPI_ISL_769281|20  ···················· ···················· ···················· ···················· ···················· ····················
AB  5277 hCoV-19/England/QEUH-C5BAA2/2020|EPI_ISL_769282|20  ···················· ···················· ···················· ···················· ······T············· ····················
AB  5278 hCoV-19/England/MILK-C60B98/2020|EPI_ISL_769283|20  ···················· ···················· ···················· ···················· ···················· ····················
AB  5279 hCoV-19/England/QEUH-C5BB36/2020|EPI_ISL_769284|20  ···················· ···················· ···················· ···················· ···················· ····················
AB  5280 hCoV-19/England/QEUH-C5BFB2/2020|EPI_ISL_769288|20  ···················· ···················· ···················· ···················· ···················· ····················
AB  5281 hCoV-19/England/QEUH-C5C40D/2020|EPI_ISL_769289|20  ···················· ···················· ···················· ···················· ···················· ····················
AB  5282 hCoV-19/England/QEUH-C5D606/2020|EPI_ISL_769290|20  ···················· ···················· ···················· ···················· ···················· ····················
AB  5283 hCoV-19/England/QEUH-C5BC15/2020|EPI_ISL_769291|20  ···················· ···················· ···················· ···················· ···················· ····················
AB  5284 hCoV-19/England/MILK-C60C4A/2020|EPI_ISL_769293|20  ···················· ···················· ···················· ···················· ···················· ····················
AB  5285 hCoV-19/England/MILK-C60DED/2020|EPI_ISL_769295|20  ···················· ···················· ···················· ···················· ···················· ····················
AB  5286 hCoV-19/England/MILK-C5FFAF/2020|EPI_ISL_769299|20  ···················· ···················· ···················· ···················· ···················· ····················
AB  5287 hCoV-19/Bangladesh/DNAS_DRICM_isl_MLS-5/2020|EPI_I  ···················· ···················· ···················· ···················· ···················· ····················
AB  5288 hCoV-19/Bangladesh/DNAS_DRICM_isl_UMCH-1/2020|EPI_  ···················· ···················· ···················· ···················· ···················· ····················
AB  5289 hCoV-19/Bangladesh/DNAS_DRICM_isl_18974/2020|EPI_I  ···················· ···················· ···················· ···················· ···················· ····················
AB  5290 hCoV-19/Bangladesh/DNAS_DRICM_isl_R-28/2020|EPI_IS  ···················· ···················· ···················· ···················· ···················· ····················
AB  5291 hCoV-19/Bangladesh/DNAS_DRICM_isl_R-2/2020|EPI_ISL  ···················· ···················· ···················· ···················· ···················· ····················
AB  5292 hCoV-19/Bangladesh/DNAS_DRICM_isl_R-6/2020|EPI_ISL  ···················· ···················· ···················· ···················· ···················· ····················
AB  5293 hCoV-19/Bangladesh/DNAS_DRICM_isl_R-3/2020|EPI_ISL  ···················· ···················· ···················· ···················· ···················· ····················
AB  5294 hCoV-19/Bangladesh/DNAS_DRICM_isl_Reza/2020|EPI_IS  ···················· ···················· ···················· ···················· ···················· ····················
AB  5295 hCoV-19/Bangladesh/GRMDRC-2/2020|EPI_ISL_775019|20  ···················· ···················· ···················· ···················· ···················· ····················
AB  5296 hCoV-19/Bangladesh/GRMDRC-3/2020|EPI_ISL_775020|20  ···················· ···················· ···················· ···················· ···················· ····················
AB  5297 hCoV-19/Canada/ON-PHL-20-03810/2020|EPI_ISL_775021  ···················· ···················· ···················· ···················· ···················· ····················
AB  5298 hCoV-19/Canada/ON-PHL-20-03811/2020|EPI_ISL_775022  ···················· ···················· ···················· ···················· ···················· ····················
AB  5300 hCoV-19/Canada/ON-PHL-20-03813/2020|EPI_ISL_775024  ···················· ·A·················· ···················· ···················· ···················· ···A················
AB  5301 hCoV-19/Canada/ON-PHL-20-03814/2020|EPI_ISL_775025  ···················· ·A·················· ···················· ···················· ···················· ···A················
AB  5302 hCoV-19/Canada/ON-PHL-20-03815/2020|EPI_ISL_775026  ···················· ···················· ···················· ···················· ···················· ····················
AB  5303 hCoV-19/Canada/ON-PHL-20-03816/2020|EPI_ISL_775027  ···················· ·A·················· ···················· ···················· ···················· ···A················
AB  5304 hCoV-19/Canada/ON-PHL-20-03817/2020|EPI_ISL_775028  ···················· ·A·················· ···················· ···················· ···················· ···A················
AB  5305 hCoV-19/Canada/ON-PHL-20-03819/2020|EPI_ISL_775029  ···················· ·A·················· ···················· ···················· ···················· ···A················
AB  5306 hCoV-19/Canada/ON-PHL-20-03820/2020|EPI_ISL_775030  ···················· ·A·················· ···················· ···················· ···················· ···A················
AB  5307 hCoV-19/Canada/ON-PHL-20-03821/2020|EPI_ISL_775031  ···················· ···················· ···················· ···················· ···················· ····················
AB  5308 hCoV-19/Canada/ON-PHL-20-03823/2020|EPI_ISL_775032  ···················· ···················· ···················· ···················· ···················· ····················
AB  5309 hCoV-19/Canada/ON-PHL-20-03833/2020|EPI_ISL_775033  ···················· ·A·················· ···················· ···················· ···················· ···A················
AB  5310 hCoV-19/Canada/ON-PHL-20-03834/2020|EPI_ISL_775034  ···················· ·A·················· ···················· ···················· ···················· ···A················
AB  5311 hCoV-19/Canada/ON-PHL-20-03751/2020|EPI_ISL_775035  ···················· ·A·················· ···················· ···················· ···················· ···A················
AB  5312 hCoV-19/Canada/ON-PHL-20-03752/2020|EPI_ISL_775036  ···················· ···················· ···················· ···················· ···················· ····················
AB  5313 hCoV-19/Canada/ON-PHL-20-03754/2020|EPI_ISL_775037  ···················· ···················· ···················· ···················· ···················· ····················
AB  5314 hCoV-19/Canada/ON-PHL-20-03755/2020|EPI_ISL_775038  ···················· ·A·················· ···················· ···················· ···················· ···A················
AB  5315 hCoV-19/Canada/ON-PHL-20-03771/2020|EPI_ISL_775080  ···················· ·········A·········· ···················· ···················· ···················· ····················
AB  5316 hCoV-19/Canada/ON-PHL-20-03772/2020|EPI_ISL_775081  ···················· ·A·················· ···················· ···················· ···················· ···A················
```

| | | | | | | | | |
|---|---|---|---|---|---|---|---|---|
| AB | 5317 | hCoV-19/Canada/ON-PHL-20-03773/2020\|EPI_ISL_775082 | ···················· | ·A·················· | ···················· | ···················· | ···················· | ···A················ |
| AB | 5318 | hCoV-19/Canada/ON-PHL-20-03774/2020\|EPI_ISL_775083 | ···················· | ·A·················· | ···················· | ···················· | ···················· | ···A················ |
| AB | 5319 | hCoV-19/Canada/ON-PHL-20-03775/2020\|EPI_ISL_775084 | ···················· | ·A·················· | ···················· | ···················· | ···················· | ···A················ |
| AB | 5320 | hCoV-19/Canada/ON-PHL-20-03776/2020\|EPI_ISL_775085 | ···················· | ···················· | ···················· | ···················· | ···················· | ···················· |
| AB | 5322 | hCoV-19/Canada/ON-PHL-20-03779/2020\|EPI_ISL_775087 | ···················· | ···················· | ···················· | ···················· | ···················· | ···················· |
| AB | 5323 | hCoV-19/Canada/ON-PHL-20-03780/2020\|EPI_ISL_775088 | ···················· | ·A·················· | ···················· | ···················· | ···················· | ···A················ |
| AB | 5324 | hCoV-19/Canada/ON-PHL-20-03781/2020\|EPI_ISL_775089 | ···················· | ·A·················· | ···················· | ···················· | ···················· | ···A················ |
| AB | 5325 | hCoV-19/Canada/ON-PHL-20-03651/2020\|EPI_ISL_775090 | ···················· | ···················· | ···················· | ···················· | ···················· | ···················· |
| AB | 5326 | hCoV-19/Canada/ON-PHL-20-03652/2020\|EPI_ISL_775091 | ···················· | ·A·················· | ···················· | ···················· | ···················· | ···A················ |
| AB | 5327 | hCoV-19/Canada/ON-PHL-20-03653/2020\|EPI_ISL_775092 | ···················· | ·A·················· | ···················· | ···················· | ···················· | ···A················ |
| AB | 5328 | hCoV-19/Canada/ON-PHL-20-03654/2020\|EPI_ISL_775093 | ···················· | ·A·················· | ···················· | ···················· | ···················· | ···A················ |
| AB | 5329 | hCoV-19/Canada/ON-PHL-20-03655/2020\|EPI_ISL_775094 | ···················· | ···················· | ···················· | ···················· | ···················· | ···················· |
| AB | 5330 | hCoV-19/Canada/ON-PHL-20-03656/2020\|EPI_ISL_775095 | ···················· | ···················· | ···················· | ···················· | ···················· | ···················· |
| AB | 5331 | hCoV-19/Canada/ON-PHL-20-03658/2020\|EPI_ISL_775096 | ···················· | ···················· | ···················· | ···················· | ···················· | ···················· |
| AB | 5332 | hCoV-19/Canada/ON-PHL-20-03659/2020\|EPI_ISL_775097 | ···················· | ·A·················· | ···················· | ···················· | ···················· | ···A················ |
| AB | 5333 | hCoV-19/Canada/ON-PHL-20-03660/2020\|EPI_ISL_775098 | ···················· | ···················· | ···················· | ···················· | ···················· | ···················· |
| AB | 5334 | hCoV-19/Canada/ON-PHL-20-03661/2020\|EPI_ISL_775099 | ···················· | ·A·················· | ···················· | ···················· | ···················· | ···A················ |
| AB | 5335 | hCoV-19/Germany/HH-hpi-p2028/2020\|EPI_ISL_776330\|2 | ···················· | ···················· | ···················· | ···················· | ···················· | ···················· |
| AB | 5336 | hCoV-19/Germany/HH-hpi-p2030/2020\|EPI_ISL_776331\|2 | ···················· | ···················· | ···················· | ···················· | ···················· | ···················· |
| AB | 5337 | hCoV-19/Germany/HH-hpi-p2039/2020\|EPI_ISL_776332\|2 | ···················· | ···················· | ···················· | ···················· | ···················· | ···················· |
| AB | 5338 | hCoV-19/Germany/HH-hpi-p2040/2020\|EPI_ISL_776333\|2 | ···················· | ···················· | ···················· | ···················· | ···················· | ···················· |
| AB | 5339 | hCoV-19/Germany/HH-hpi-p2041/2020\|EPI_ISL_776334\|2 | ···················· | ·A·················· | ···················· | ···················· | ···················· | ···················· |
| AB | 5340 | hCoV-19/Germany/HH-hpi-p2043/2020\|EPI_ISL_776335\|2 | ···················· | ···················· | ···················· | ···················· | ···················· | ···················· |
| AB | 5341 | hCoV-19/Germany/HH-hpi-p2044/2020\|EPI_ISL_776336\|2 | ···················· | ···················· | ···················· | ···················· | ···················· | ···················· |
| AB | 5342 | hCoV-19/Germany/HH-hpi-p2045/2020\|EPI_ISL_776337\|2 | ···················· | ···················· | ···················· | ···················· | ···················· | ···················· |
| AB | 5343 | hCoV-19/Germany/HH-hpi-p2080/2020\|EPI_ISL_776349\|2 | ···················· | ···················· | ···················· | ···················· | ···················· | ···················· |
| AB | 5344 | hCoV-19/Germany/HH-hpi-p2087/2020\|EPI_ISL_776350\|2 | ···················· | ···················· | ···················· | ···················· | ···················· | ···················· |
| AB | 5345 | hCoV-19/Germany/HH-hpi-p2104/2020\|EPI_ISL_776351\|2 | ···················· | ···················· | ···················· | ···················· | ···················· | ···················· |
| AB | 5346 | hCoV-19/Germany/HH-hpi-p2105/2020\|EPI_ISL_776352\|2 | ···················· | ···················· | ···················· | ···················· | ···················· | ···················· |
| AB | 5347 | hCoV-19/Germany/HH-hpi-p2107/2020\|EPI_ISL_776353\|2 | ···················· | ···················· | ···················· | ···················· | ···················· | ···················· |
| AB | 5348 | hCoV-19/Germany/HH-hpi-p2109/2020\|EPI_ISL_776354\|2 | ···················· | ···················· | ···················· | ···················· | ···················· | ···················· |
| AB | 5349 | hCoV-19/Germany/HH-hpi-p2110/2020\|EPI_ISL_776355\|2 | ···················· | ···················· | ···················· | ···················· | ···················· | ···················· |
| AB | 5350 | hCoV-19/Germany/HH-hpi-p2111/2020\|EPI_ISL_776356\|2 | ···················· | ···················· | ···················· | ···················· | ···················· | ···················· |
| AB | 5351 | hCoV-19/Germany/HH-hpi-p2113/2020\|EPI_ISL_776357\|2 | ···················· | ···················· | ···················· | ···················· | ···················· | ···················· |
| AB | 5352 | hCoV-19/Germany/HH-hpi-p2114/2020\|EPI_ISL_776358\|2 | ···················· | ···················· | ···················· | ···················· | ···················· | ···················· |
| AB | 5353 | hCoV-19/Germany/HH-hpi-p2115/2020\|EPI_ISL_776359\|2 | ···················· | ···················· | ···················· | ···················· | ···················· | ···················· |
| AB | 5354 | hCoV-19/Germany/HH-hpi-p2116/2020\|EPI_ISL_776360\|2 | ···················· | ···················· | ···················· | ···················· | ···················· | ···················· |
| AB | 5355 | hCoV-19/Germany/HH-hpi-p2118/2020\|EPI_ISL_776361\|2 | ···················· | ···················· | ···················· | ···················· | ···················· | ···················· |
| AB | 5356 | hCoV-19/Germany/HH-hpi-p2119/2020\|EPI_ISL_776362\|2 | ···················· | ···················· | ···················· | ···················· | ·············T······ | ···················· |
| AB | 5357 | hCoV-19/Germany/HH-hpi-p2120/2020\|EPI_ISL_776363\|2 | ···················· | ···················· | ···················· | ···················· | ···················· | ···················· |
| AB | 5358 | hCoV-19/Germany/HH-hpi-p2124/2020\|EPI_ISL_776364\|2 | ···················· | ···················· | ···················· | ···················· | ···················· | ···················· |
| AB | 5359 | hCoV-19/Germany/HH-hpi-p2125/2020\|EPI_ISL_776365\|2 | ···················· | ···················· | ···················· | ···················· | ···················· | ···················· |
| AB | 5360 | hCoV-19/Germany/HH-hpi-p2126/2020\|EPI_ISL_776366\|2 | ···················· | ···················· | ···················· | ···················· | ···················· | ···················· |
| AB | 5361 | hCoV-19/Germany/HH-hpi-p2127/2020\|EPI_ISL_776367\|2 | ···················· | ···················· | ···················· | ···················· | ···················· | ···················· |
| AB | 5362 | hCoV-19/Germany/HH-hpi-p2128/2020\|EPI_ISL_776368\|2 | ···················· | ···················· | ···················· | ···················· | ···················· | ···················· |
| AB | 5363 | hCoV-19/Germany/HH-hpi-p2129/2020\|EPI_ISL_776369\|2 | ···················· | ···················· | ···················· | ···················· | ···················· | ···················· |
| AB | 5364 | hCoV-19/England/CAMC-CB7840/2020\|EPI_ISL_777689\|20 | ···················· | ···················· | ···················· | ···················· | ···················· | ···················· |
| AB | 5365 | hCoV-19/England/CAMC-CB7B38/2020\|EPI_ISL_777696\|20 | ···················· | ···················· | ···················· | ···················· | ···················· | ···················· |
| AB | 5366 | hCoV-19/England/CAMC-CB7145/2020\|EPI_ISL_777697\|20 | ···················· | ·A·················· | ···················· | ···················· | ···················· | ···A················ |
| AB | 5367 | hCoV-19/USA/LA-EVTL1324/2020\|EPI_ISL_778990\|2020-1 | ···················· | ···················· | ···················· | ···················· | ···················· | ···················· |
| AB | 5368 | hCoV-19/USA/LA-EVTL1325/2020\|EPI_ISL_778991\|2020-1 | ···················· | ···················· | ···················· | ···················· | ···················· | ···················· |
| AB | 5369 | hCoV-19/USA/LA-EVTL1326/2020\|EPI_ISL_778992\|2020-1 | ···················· | ···················· | ···················· | ···················· | ···················· | ···················· |
| AB | 5370 | hCoV-19/USA/LA-EVTL1327/2020\|EPI_ISL_778993\|2020-1 | ···················· | ···················· | ···················· | ···················· | ···················· | ···················· |
| AB | 5371 | hCoV-19/USA/LA-EVTL1328/2020\|EPI_ISL_778994\|2020-1 | ···················· | ···················· | ···················· | ···················· | ···················· | ···················· |
| AB | 5372 | hCoV-19/USA/LA-EVTL1329/2020\|EPI_ISL_778995\|2020-1 | ···················· | ···················· | ···················· | ···················· | ···················· | ···················· |
| AB | 5373 | hCoV-19/USA/LA-EVTL1330/2020\|EPI_ISL_778996\|2020-1 | ···················· | ···················· | ···················· | ···················· | ···················· | ···················· |
| AB | 5374 | hCoV-19/USA/LA-EVTL1331/2020\|EPI_ISL_778997\|2020-1 | ···················· | ···················· | ···················· | ···················· | ···················· | ···················· |
| AB | 5375 | hCoV-19/USA/LA-EVTL1332/2020\|EPI_ISL_778998\|2020-1 | ···················· | ···················· | ···················· | ···················· | ···················· | ···················· |
| AB | 5376 | hCoV-19/USA/LA-EVTL1333/2020\|EPI_ISL_778999\|2020-1 | ···················· | ···················· | ···················· | ···················· | ···················· | ···················· |
| AB | 5377 | hCoV-19/Netherlands/LI-EMC-132/2020\|EPI_ISL_763100 | ···················· | ···················· | ···················· | ···················· | ···················· | ···················· |
| AB | 5378 | hCoV-19/Netherlands/LI-EMC-133/2020\|EPI_ISL_763101 | ···················· | ···················· | ···················· | ···················· | ···················· | ···················· |
| AB | 5379 | hCoV-19/Netherlands/LI-EMC-134/2020\|EPI_ISL_763102 | ···················· | ···················· | ···················· | ···················· | ···················· | ···················· |
| AB | 5380 | hCoV-19/Netherlands/LI-EMC-135/2020\|EPI_ISL_763103 | ···················· | ···················· | ···················· | ···················· | ···················· | ···················· |
| AB | 5381 | hCoV-19/Netherlands/LI-EMC-137/2020\|EPI_ISL_763105 | ···················· | ···················· | ···················· | ···················· | ···················· | ···················· |
| AB | 5382 | hCoV-19/Netherlands/LI-EMC-139/2020\|EPI_ISL_763107 | ···················· | ···················· | ···················· | ···················· | ···················· | ···················· |
| AB | 5383 | hCoV-19/Netherlands/LI-EMC-140/2020\|EPI_ISL_763108 | ···················· | ···················· | ···················· | ···················· | ···················· | ···················· |
| AB | 5384 | hCoV-19/Netherlands/LI-EMC-141/2020\|EPI_ISL_763109 | ···················· | ···················· | ···················· | ···················· | ···················· | ···················· |
| AB | 5385 | hCoV-19/Netherlands/LI-EMC-142/2020\|EPI_ISL_763110 | ···················· | ···················· | ···················· | ···················· | ···················· | ···················· |
| AB | 5386 | hCoV-19/Netherlands/LI-EMC-143/2020\|EPI_ISL_763111 | ···················· | ···················· | ···················· | ···················· | ···················· | ···················· |
| AB | 5387 | hCoV-19/Netherlands/LI-EMC-144/2020\|EPI_ISL_763112 | ···················· | ···················· | ···················· | ···················· | ···················· | ···················· |
| AB | 5388 | hCoV-19/Netherlands/LI-EMC-145/2020\|EPI_ISL_763113 | ············T······· | ···················· | ···················· | ···················· | ···················· | ···················· |
| AB | 5389 | hCoV-19/Netherlands/LI-EMC-146/2020\|EPI_ISL_763114 | ···················· | ···················· | ···················· | ···················· | ···················· | ···················· |
| AB | 5390 | hCoV-19/Netherlands/LI-EMC-147/2020\|EPI_ISL_763115 | ···················· | ···················· | ···················· | ···················· | ···················· | ···················· |
| AB | 5391 | hCoV-19/Netherlands/LI-EMC-148/2020\|EPI_ISL_763116 | ···················· | ···················· | ···················· | ···················· | ···················· | ···················· |
| AB | 5392 | hCoV-19/Netherlands/LI-EMC-150/2020\|EPI_ISL_763118 | ···················· | ···················· | ···················· | ···················· | ···················· | ···················· |
| AB | 5393 | hCoV-19/Netherlands/LI-EMC-151/2020\|EPI_ISL_763119 | ···················· | ···················· | ···················· | ···················· | ···················· | ···················· |
| AB | 5394 | hCoV-19/Netherlands/LI-EMC-152/2020\|EPI_ISL_763120 | ···················· | ···················· | ···················· | ···················· | ···················· | ···················· |
| AB | 5395 | hCoV-19/Netherlands/LI-EMC-153/2020\|EPI_ISL_763121 | ···················· | ···················· | ···················· | ···················· | ···················· | ···················· |
| AB | 5396 | hCoV-19/Netherlands/LI-EMC-156/2020\|EPI_ISL_763124 | ···················· | ···················· | ···················· | ···················· | ···················· | ···················· |
| AB | 5397 | hCoV-19/Netherlands/LI-EMC-157/2020\|EPI_ISL_763125 | ···················· | ···················· | ···················· | ···················· | ···················· | ···················· |

| | | | | | | | | |
|---|---|---|---|---|---|---|---|---|
| AB | 5398 hCoV-19/Netherlands/LI-EMC-158/2020\|EPI_ISL_763126 | .................... | .................... | ..................... | .................... | .................... | .................... | |
| AB | 5399 hCoV-19/Netherlands/LI-EMC-159/2020\|EPI_ISL_763127 | .................... | .................... | ..................... | .................... | .................... | .................... | |
| AB | 5400 hCoV-19/Netherlands/LI-EMC-160/2020\|EPI_ISL_763128 | .................... | .................... | ..................... | .................... | .................... | .................... | |
| AB | 5401 hCoV-19/Netherlands/LI-EMC-161/2020\|EPI_ISL_763129 | .................... | .................... | ..................... | .................... | .................... | .................... | |
| AB | 5402 hCoV-19/Netherlands/NB-EMC-472/2020\|EPI_ISL_763130 | .................... | .................... | ..................... | .................... | .................... | .................... | |
| AB | 5403 hCoV-19/Netherlands/NB-EMC-473/2020\|EPI_ISL_763131 | .................... | .................... | ..................... | .................... | .................... | .................... | |
| AB | 5404 hCoV-19/Netherlands/NB-EMC-474/2020\|EPI_ISL_763132 | .................... | .................... | ..................... | .................... | .................... | .................... | |
| AB | 5405 hCoV-19/Netherlands/NB-EMC-475/2020\|EPI_ISL_763133 | .................... | .................... | ..................... | .................... | .................... | .................... | |
| AB | 5406 hCoV-19/Netherlands/NB-EMC-476/2020\|EPI_ISL_763134 | .................... | .................... | ..................... | .................... | .................... | .................... | |
| AB | 5407 hCoV-19/Netherlands/NB-EMC-477/2020\|EPI_ISL_763135 | .................... | .................... | ..................... | .................... | .................... | .................... | |
| AB | 5408 hCoV-19/Netherlands/NB-EMC-478/2020\|EPI_ISL_763136 | .................... | .................... | ..................... | .................... | .................... | .................... | |
| AB | 5409 hCoV-19/Netherlands/NB-EMC-479/2020\|EPI_ISL_763137 | .................... | .................... | ..................... | .................... | .................... | .................... | |
| AB | 5411 hCoV-19/Northern Ireland/NIRE-234EBC/2020\|EPI_ISL_ | .................... | .................... | ..................... | .................... | .................... | .................... | |
| AB | 5412 hCoV-19/Northern Ireland/NIRE-234F8C/2020\|EPI_ISL_ | .................... | .................... | ..................... | .................... | .................... | .................... | |
| AB | 5413 hCoV-19/Northern Ireland/NIRE-234FD7/2020\|EPI_ISL_ | .................... | .................... | ..................... | .................... | .................... | .................... | |
| AB | 5414 hCoV-19/Northern Ireland/NIRE-234FF5/2020\|EPI_ISL_ | .................... | .................... | ..................... | .................... | .................... | .................... | |
| AB | 5417 hCoV-19/Northern Ireland/NIRE-235097/2020\|EPI_ISL_ | .................... | .................... | ..................... | .................... | .................... | .................... | |
| AB | 5418 hCoV-19/Northern Ireland/NIRE-2350A6/2020\|EPI_ISL_ | .................... | .................... | ..................... | .................... | .................... | .................... | |
| AB | 5419 hCoV-19/Northern Ireland/NIRE-2350B5/2020\|EPI_ISL_ | .................... | .................... | ..................... | .................... | .................... | .................... | |
| AB | 5420 hCoV-19/Northern Ireland/NIRE-2350D3/2020\|EPI_ISL_ | .................... | .................... | ..................... | .................... | .................... | .................... | |
| AB | 5421 hCoV-19/Northern Ireland/NIRE-2350E2/2020\|EPI_ISL_ | .................... | .................... | ..................... | .................... | NNN................. | .................... | 1 |
| AB | 5422 hCoV-19/Northern Ireland/NIRE-2350F1/2020\|EPI_ISL_ | .................... | .................... | ..................... | .................... | .................... | .................... | |
| AB | 5423 hCoV-19/Northern Ireland/NIRE-23510D/2020\|EPI_ISL_ | .................... | .................... | ..................... | .................... | .................... | .................... | |
| AB | 5424 hCoV-19/Northern Ireland/NIRE-2352A0/2020\|EPI_ISL_ | .................... | .................... | ..................... | .................... | .................... | .................... | |
| AB | 5425 hCoV-19/Northern Ireland/NIRE-235307/2020\|EPI_ISL_ | .................... | .................... | ..................... | .................... | .................... | .................... | |
| AB | 5427 hCoV-19/Northern Ireland/NIRE-235343/2020\|EPI_ISL_ | .................... | .................... | ..................... | .................... | .................... | .................... | |
| AB | 5428 hCoV-19/Northern Ireland/NIRE-235352/2020\|EPI_ISL_ | .................... | .................... | ..................... | .................... | .................... | .................... | |
| AB | 5429 hCoV-19/Northern Ireland/NIRE-235370/2020\|EPI_ISL_ | .................... | .................... | ..................... | .................... | .................... | .................... | |
| AB | 5430 hCoV-19/Northern Ireland/NIRE-2353AD/2020\|EPI_ISL_ | .................... | .................... | ..................... | .................... | .................... | .................... | |
| AB | 5432 hCoV-19/Northern Ireland/NIRE-23545F/2020\|EPI_ISL_ | .................... | .................... | ..................... | .................... | .NN................. | .................... | |
| AB | 5434 hCoV-19/Northern Ireland/NIRE-23549B/2020\|EPI_ISL_ | .................... | .................... | ..................... | .................... | .................... | .................... | |
| AB | 5435 hCoV-19/Northern Ireland/NIRE-2354AA/2020\|EPI_ISL_ | .................... | .................... | ..................... | .................... | .................... | .................... | |
| AB | 5436 hCoV-19/Northern Ireland/NIRE-2354D7/2020\|EPI_ISL_ | .................... | .................... | ..................... | .................... | .................... | .................... | |
| AB | 5437 hCoV-19/Northern Ireland/NIRE-2354E6/2020\|EPI_ISL_ | .................... | .................... | ..................... | .................... | .................... | .................... | |
| AB | 5439 hCoV-19/Northern Ireland/NIRE-23553E/2020\|EPI_ISL_ | .................... | .................... | ..................... | .................... | .................... | .................... | |
| AB | 5440 hCoV-19/Northern Ireland/NIRE-23555C/2020\|EPI_ISL_ | .................... | .................... | ..................... | .................... | .................... | .................... | |
| AB | 5441 hCoV-19/Northern Ireland/NIRE-23556B/2020\|EPI_ISL_ | .................... | .................... | ..................... | .................... | .................... | .................... | |
| AB | 5442 hCoV-19/Northern Ireland/NIRE-23557A/2020\|EPI_ISL_ | .................... | .................... | ..................... | .................... | .................... | .................... | |
| AB | 5443 hCoV-19/Northern Ireland/NIRE-235589/2020\|EPI_ISL_ | .................... | .................... | ..................... | .................... | .................... | .................... | |
| AB | 5444 hCoV-19/England/NORW-220F74/2020\|EPI_ISL_764445\|20 | .................... | .................... | ..................... | .................... | .................... | .................... | |
| AB | 5445 hCoV-19/England/NORW-221113/2020\|EPI_ISL_764446\|20 | .................... | .................... | ..................... | .................... | .................... | .................... | |
| AB | 5446 hCoV-19/England/NORW-22115F/2020\|EPI_ISL_764447\|20 | .................... | .................... | ..................... | .................... | .................... | .................... | |
| AB | 5447 hCoV-19/England/NORW-22118C/2020\|EPI_ISL_764448\|20 | .................... | .................... | ..................... | .................... | .................... | .................... | |
| AB | 5448 hCoV-19/England/NORW-2211C8/2020\|EPI_ISL_764449\|20 | .................... | .................... | ..................... | .................... | .................... | .................... | |
| AB | 5449 hCoV-19/England/NORW-2211E6/2020\|EPI_ISL_764450\|20 | .................... | .................... | ..................... | .................... | .................... | .................... | |
| AB | 5450 hCoV-19/England/NORW-221210/2020\|EPI_ISL_764451\|20 | .................... | .................... | ..................... | .................... | .................... | .................... | |
| AB | 5451 hCoV-19/England/NORW-22122F/2020\|EPI_ISL_764452\|20 | .................... | .................... | ..................... | .................... | .................... | .................... | |
| AB | 5452 hCoV-19/England/NORW-22123E/2020\|EPI_ISL_764453\|20 | .................... | .................... | ..................... | .................... | .................... | .................... | |
| AB | 5453 hCoV-19/England/NORW-22124D/2020\|EPI_ISL_764454\|20 | .................... | .................... | ..................... | .................... | .................... | .................... | |
| AB | 5454 hCoV-19/England/NORW-22190C/2020\|EPI_ISL_764455\|20 | .................... | .................... | ..................... | .................... | .................... | .................... | |
| AB | 5455 hCoV-19/England/NORW-221948/2020\|EPI_ISL_764456\|20 | .................... | .................... | ..................... | .................... | .................... | .................... | |
| AB | 5456 hCoV-19/England/NORW-221975/2020\|EPI_ISL_764457\|20 | .................... | .................... | ..................... | .................... | .................... | .................... | |
| AB | 5457 hCoV-19/England/NORW-2219A2/2020\|EPI_ISL_764458\|20 | .................... | .................... | ..................... | .................... | .................... | .................... | |
| AB | 5458 hCoV-19/England/NORW-2219DF/2020\|EPI_ISL_764459\|20 | .................... | .................... | ..................... | .................... | .................... | .................... | |
| AB | 5459 hCoV-19/England/NORW-2219FD/2020\|EPI_ISL_764460\|20 | .................... | .................... | ..................... | .................... | .................... | .................... | |
| AB | 5460 hCoV-19/England/NORW-221A18/2020\|EPI_ISL_764461\|20 | .................... | .................... | ..................... | .................... | .................... | .................... | |
| AB | 5461 hCoV-19/England/NORW-221A36/2020\|EPI_ISL_764462\|20 | .................... | .................... | ..................... | .................... | .................... | .................... | |
| AB | 5462 hCoV-19/England/NORW-221A63/2020\|EPI_ISL_764463\|20 | .................... | .................... | ..................... | .................... | .................... | .................... | |
| AB | 5463 hCoV-19/England/NORW-221ABE/2020\|EPI_ISL_764464\|20 | .................... | .................... | ..................... | .................... | .................... | .................... | |
| AB | 5464 hCoV-19/England/NORW-221AEB/2020\|EPI_ISL_764465\|20 | .................... | .................... | ..................... | .................... | .................... | .................... | |
| AB | 5465 hCoV-19/England/NORW-221B06/2020\|EPI_ISL_764466\|20 | .................... | .................... | ..................... | .................... | .................... | .................... | |
| AB | 5466 hCoV-19/England/NORW-221B60/2020\|EPI_ISL_764467\|20 | .................... | .................... | ..................... | .................... | .................... | .................... | |
| AB | 5467 hCoV-19/England/NORW-221BAC/2020\|EPI_ISL_764468\|20 | .................... | .................... | ..................... | .................... | .................... | .................... | |
| AB | 5468 hCoV-19/USA/MA-MGH-03082/2020\|EPI_ISL_765701\|2020- | .................... | .................... | ..................... | .................... | .................... | .................... | |
| AB | 5469 hCoV-19/USA/MA-MGH-03083/2020\|EPI_ISL_765702\|2020- | .................... | .................... | ..................... | .................... | .................... | .................... | |
| AB | 5470 hCoV-19/USA/MA-MGH-03085/2020\|EPI_ISL_765703\|2020- | .................... | .................... | ..................... | .................... | .................... | .................... | |
| AB | 5471 hCoV-19/USA/MA-MGH-03101/2020\|EPI_ISL_765704\|2020- | .................... | .................... | ..................... | .................... | .................... | .................... | |
| AB | 5472 hCoV-19/USA/MA-MGH-03110/2020\|EPI_ISL_765705\|2020- | .................... | .................... | ..................... | .................... | .................... | .................... | |
| AB | 5473 hCoV-19/USA/MA-MGH-03111/2020\|EPI_ISL_765706\|2020- | .................... | .................... | ..................... | .................... | .................... | .................... | |
| AB | 5474 hCoV-19/USA/MA-MGH-03116/2020\|EPI_ISL_765708\|2020- | .................... | .................... | ..................... | .................... | .................... | .................... | |
| AB | 5475 hCoV-19/USA/MA-MGH-03117/2020\|EPI_ISL_765709\|2020- | .................... | .A.................. | ..................... | .................... | .................... | .................... | |
| AB | 5476 hCoV-19/USA/MA-MGH-03125/2020\|EPI_ISL_765712\|2020- | .................... | .................... | ..................... | .................... | .................... | .................... | |
| AB | 5477 hCoV-19/USA/MA-MGH-03128/2020\|EPI_ISL_765713\|2020- | .................... | .................... | ..................... | .................... | .................... | .................... | |
| AB | 5478 hCoV-19/USA/MA-MGH-03139/2020\|EPI_ISL_765714\|2020- | .................... | .................... | ..................... | .................... | .................... | .................... | |
| AB | 5479 hCoV-19/USA/MA-MGH-03141/2020\|EPI_ISL_765715\|2020- | .................... | .................... | ..................... | .................... | .................... | .................... | |
| AB | 5480 hCoV-19/USA/MA-MGH-03144/2020\|EPI_ISL_765716\|2020- | .................... | .................... | ..................... | .................... | .................... | .................... | |
| AB | 5481 hCoV-19/USA/MA-MGH-03145/2020\|EPI_ISL_765717\|2020- | .................... | .................... | ..................... | .................... | .................... | .................... | |
| AB | 5482 hCoV-19/USA/MA-MGH-03146/2020\|EPI_ISL_765718\|2020- | .................... | .................... | ..................... | .................... | .................... | .................... | |
| AB | 5483 hCoV-19/USA/MA-MGH-03149/2020\|EPI_ISL_765720\|2020- | .................... | .................... | ..................... | .................... | .................... | .................... | |
| AB | 5484 hCoV-19/USA/MA-MGH-03150/2020\|EPI_ISL_765721\|2020- | .................... | .................... | ..................... | .................... | .................... | .................... | |

| | | | | | | | |
|---|---|---|---|---|---|---|---|
| AB | 5485 hCoV-19/USA/MA-MGH-03155/2020\|EPI_ISL_765722\|2020- | ···················· | ···················· | ····················· | ···················· | ···················· | ···················· |
| AB | 5486 hCoV-19/USA/MA-MGH-03156/2020\|EPI_ISL_765723\|2020- | ···················· | ···················· | ····················· | ···················· | ···················· | ···················· |
| AB | 5487 hCoV-19/USA/MA-MGH-03161/2020\|EPI_ISL_765724\|2020- | ···················· | ···················· | ····················· | ···················· | ···················· | ···················· |
| AB | 5488 hCoV-19/USA/MA-MGH-03162/2020\|EPI_ISL_765725\|2020- | ···················· | ····A··············· | ····················· | ···················· | ···················· | ···················· |
| AB | 5489 hCoV-19/USA/MA-MGH-03165/2020\|EPI_ISL_765726\|2020- | ···················· | ···················· | ····················· | ···················· | ···················· | ···················· |
| AB | 5490 hCoV-19/USA/MA-MGH-03167/2020\|EPI_ISL_765727\|2020- | ···················· | ···················· | ····················· | ···················· | ···················· | ···················· |
| AB | 5491 hCoV-19/USA/MA-MGH-03168/2020\|EPI_ISL_765728\|2020- | ···················· | ···················· | ····················· | ···················· | ···················· | ···················· |
| AB | 5492 hCoV-19/USA/MA-MGH-03172/2020\|EPI_ISL_765729\|2020- | ···················· | ···················· | ····················· | ···················· | ···················· | ···················· |
| AB | 5493 hCoV-19/USA/MA-MGH-03173/2020\|EPI_ISL_765730\|2020- | ···················· | ···················· | ····················· | ···················· | ···················· | ···················· |
| AB | 5494 hCoV-19/USA/MA-MGH-03175/2020\|EPI_ISL_765732\|2020- | ···················· | ···················· | ····················· | ···················· | ···················· | ···················· |
| AB | 5495 hCoV-19/USA/MA-MGH-03176/2020\|EPI_ISL_765733\|2020- | ···················· | ···················· | ····················· | ···················· | ···················· | ···················· |
| AB | 5496 hCoV-19/USA/MA-MGH-03177/2020\|EPI_ISL_765734\|2020- | ···················· | ···················· | ····················· | ···················· | ···················· | ···················· |
| AB | 5497 hCoV-19/USA/MA-MGH-03178/2020\|EPI_ISL_765735\|2020- | ···················· | ·A·················· | ····················· | ···················· | ···················· | ···················· |
| AB | 5498 hCoV-19/USA/MA-MGH-03179/2020\|EPI_ISL_765736\|2020- | ···················· | ···················· | ····················· | ···················· | ···················· | ···················· |
| AB | 5499 hCoV-19/USA/MA-MGH-03182/2020\|EPI_ISL_765737\|2020- | ···················· | ···················· | ····················· | ···················· | ···················· | ···················· |
| AB | 5500 hCoV-19/USA/MA-MGH-03183/2020\|EPI_ISL_765738\|2020- | ···················· | ···················· | ····················· | ···················· | ···················· | ···················· |
| AB | 5501 hCoV-19/USA/MA-MGH-03184/2020\|EPI_ISL_765739\|2020- | ···················· | ···················· | ····················· | ···················· | ···················· | ···················· |
| AB | 5502 hCoV-19/USA/MA-MGH-03188/2020\|EPI_ISL_765740\|2020- | ···················· | ···················· | ····················· | ···················· | ···················· | ···················· |
| AB | 5503 hCoV-19/USA/MA-MGH-03190/2020\|EPI_ISL_765741\|2020- | ···················· | ···················· | ····················· | ···················· | ···················· | ···················· |
| AB | 5504 hCoV-19/USA/MA-MGH-03191/2020\|EPI_ISL_765742\|2020- | ···················· | ···················· | ····················· | ···················· | ···················· | ···················· |
| AB | 5505 hCoV-19/USA/MA-MGH-03192/2020\|EPI_ISL_765743\|2020- | ···················· | ···················· | ····················· | ···················· | ···················· | ···················· |
| AB | 5506 hCoV-19/USA/MA-MGH-03202/2020\|EPI_ISL_765744\|2020- | ···················· | ···················· | ····················· | ···················· | ···················· | ···················· |
| AB | 5507 hCoV-19/USA/MA-MGH-03244/2020\|EPI_ISL_765756\|2020- | ···················· | ···················· | ····················· | ···················· | ···················· | ···················· |
| AB | 5508 hCoV-19/USA/MA-MGH-03247/2020\|EPI_ISL_765757\|2020- | ···················· | ···················· | ····················· | ···················· | ···················· | ···················· |
| AB | 5509 hCoV-19/USA/MA-MGH-03248/2020\|EPI_ISL_765758\|2020- | ···················· | ···················· | ····················· | ···················· | ······T············· | ···················· |
| AB | 5510 hCoV-19/USA/MA-MGH-03251/2020\|EPI_ISL_765759\|2020- | ···················· | ···················· | ····················· | ···················· | ···················· | ···················· |
| AB | 5511 hCoV-19/USA/MA-MGH-03253/2020\|EPI_ISL_765760\|2020- | ···················· | ···················· | ····················· | ···················· | ···················· | ···················· |
| AB | 5512 hCoV-19/USA/MA-MGH-03257/2020\|EPI_ISL_765761\|2020- | ···················· | ·A·················· | ····················· | ···················· | ···················· | ···················· |
| AB | 5513 hCoV-19/USA/MA-MGH-03261/2020\|EPI_ISL_765762\|2020- | ···················· | ···················· | ····················· | ···················· | ···················· | ···················· |
| AB | 5514 hCoV-19/USA/MA-MGH-03263/2020\|EPI_ISL_765763\|2020- | ···················· | ···················· | ····················· | ···················· | ···················· | ···················· |
| AB | 5515 hCoV-19/USA/MA-MGH-03266/2020\|EPI_ISL_765764\|2020- | ···················· | ···················· | ····················· | ···················· | ···················· | ···················· |
| AB | 5516 hCoV-19/USA/MA-MGH-03268/2020\|EPI_ISL_765765\|2020- | ···················· | ···················· | ····················· | ···················· | ···················· | ···················· |
| AB | 5517 hCoV-19/USA/MA-MGH-03269/2020\|EPI_ISL_765766\|2020- | ···················· | ···················· | ····················· | ···················· | ···················· | ···················· |
| AB | 5518 hCoV-19/USA/MA-MGH-03271/2020\|EPI_ISL_765767\|2020- | ···················· | ···················· | ····················· | ···················· | ···················· | ···················· |
| AB | 5519 hCoV-19/USA/MA-MGH-03274/2020\|EPI_ISL_765768\|2020- | ···················· | ·A·················· | ····················· | ···················· | ···················· | ···················· |
| AB | 5520 hCoV-19/USA/MA-MGH-03277/2020\|EPI_ISL_765769\|2020- | ···················· | ···················· | ····················· | ···················· | ···················· | ···················· |
| AB | 5521 hCoV-19/USA/MA-MGH-03282/2020\|EPI_ISL_765771\|2020- | ···················· | ·A·················· | ····················· | ···················· | ···················· | ···················· |
| AB | 5522 hCoV-19/USA/MA-MGH-03283/2020\|EPI_ISL_765772\|2020- | ···················· | ···················· | ····················· | ···················· | ···················· | ···················· |
| AB | 5523 hCoV-19/USA/MA-MGH-03287/2020\|EPI_ISL_765773\|2020- | ···················· | ···················· | ····················· | ···················· | ···················· | ···················· |
| AB | 5524 hCoV-19/USA/MA-MGH-03288/2020\|EPI_ISL_765774\|2020- | ···················· | ···················· | ····················· | ···················· | ···················· | ···················· |
| AB | 5525 hCoV-19/USA/MA-MGH-03294/2020\|EPI_ISL_765775\|2020- | ···················· | ···················· | ····················· | ···················· | ···················· | ···················· |
| AB | 5526 hCoV-19/USA/MA-MGH-03296/2020\|EPI_ISL_765776\|2020- | ···················· | ···················· | ····················· | ···················· | ···················· | ···················· |
| AB | 5528 hCoV-19/USA/MA-MGH-03307/2020\|EPI_ISL_765778\|2020- | ···················· | ···················· | ····················· | ···················· | ···················· | ···················· |
| AB | 5529 hCoV-19/USA/MA-MGH-03308/2020\|EPI_ISL_765779\|2020- | ···················· | ···················· | ····················· | ···················· | ···················· | ···················· |
| AB | 5530 hCoV-19/USA/MA-MGH-03309/2020\|EPI_ISL_765780\|2020- | ···················· | ···················· | ····················· | ···················· | ···················· | ···················· |
| AB | 5531 hCoV-19/USA/MA-MGH-03310/2020\|EPI_ISL_765781\|2020- | ···················· | ·A·················· | ····················· | ···················· | ···················· | ···················· |
| AB | 5532 hCoV-19/USA/MA-MGH-03314/2020\|EPI_ISL_765782\|2020- | ···················· | ···················· | ····················· | ···················· | ···················· | ···················· |
| AB | 5533 hCoV-19/USA/MA-MGH-03316/2020\|EPI_ISL_765783\|2020- | ···················· | ···················· | ····················· | ···················· | ···················· | ···················· |
| AB | 5534 hCoV-19/USA/MA-MGH-03318/2020\|EPI_ISL_765785\|2020- | ···················· | ···················· | ····················· | ···················· | ···················· | ···················· |
| AB | 5535 hCoV-19/USA/MA-MGH-03319/2020\|EPI_ISL_765786\|2020- | ···················· | ···················· | ····················· | ···················· | ···················· | ···················· |
| AB | 5536 hCoV-19/USA/MA-MGH-03324/2020\|EPI_ISL_765787\|2020- | ···················· | ···················· | ····················· | ···················· | ···················· | ···················· |
| AB | 5537 hCoV-19/USA/MA-MGH-03330/2020\|EPI_ISL_765788\|2020- | ···················· | ···················· | ····················· | ···················· | ···················· | ···················· |
| AB | 5538 hCoV-19/USA/MA-MGH-03331/2020\|EPI_ISL_765789\|2020- | ···················· | ···················· | ····················· | ···················· | ···················· | ···················· |
| AB | 5539 hCoV-19/USA/MA-MGH-03332/2020\|EPI_ISL_765790\|2020- | ···················· | ···················· | ····················· | ···················· | ···················· | ···················· |
| AB | 5540 hCoV-19/USA/MA-MGH-03336/2020\|EPI_ISL_765791\|2020- | ···················· | ···················· | ····················· | ···················· | ···················· | ···················· |
| AB | 5541 hCoV-19/USA/MA-MGH-03337/2020\|EPI_ISL_765792\|2020- | ···················· | ···················· | ····················· | ···················· | ···················· | ···················· |
| AB | 5542 hCoV-19/USA/MA-MGH-03339/2020\|EPI_ISL_765793\|2020- | ···················· | ···················· | ····················· | ···················· | ···················· | ···················· |
| AB | 5543 hCoV-19/USA/MA-MGH-03342/2020\|EPI_ISL_765794\|2020- | ···················· | ···················· | ····················· | ···················· | ···················· | ···················· |
| AB | 5544 hCoV-19/USA/MA-MGH-03345/2020\|EPI_ISL_765795\|2020- | ···················· | ···················· | ····················· | ···················· | ···················· | ···················· |
| AB | 5545 hCoV-19/USA/MA-MGH-03346/2020\|EPI_ISL_765796\|2020- | ···················· | ···················· | ····················· | ···················· | ···················· | ···················· |
| AB | 5546 hCoV-19/USA/MA-MGH-03347/2020\|EPI_ISL_765797\|2020- | ···················· | ···················· | ····················· | ···················· | ···················· | ···················· |
| AB | 5547 hCoV-19/USA/MA-MGH-03349/2020\|EPI_ISL_765798\|2020- | ···················· | ···················· | ····················· | ···················· | ···················· | ···················· |
| AB | 5548 hCoV-19/England/QEUH-C5B94B/2020\|EPI_ISL_769246\|20 | ···················· | ···················· | ····················· | ···················· | ···················· | ···················· |
| AB | 5549 hCoV-19/England/QEUH-C5BB09/2020\|EPI_ISL_769249\|20 | ···················· | ·A·················· | ····················· | ···················· | ···················· | ···················· |
| AB | 5550 hCoV-19/England/MILK-C613EA/2020\|EPI_ISL_769251\|20 | ···················· | ···················· | ····················· | ···················· | ···················· | ···A················ |
| AB | 5551 hCoV-19/England/QEUH-C5BA66/2020\|EPI_ISL_769252\|20 | ···················· | ···················· | ····················· | ···················· | ···················· | ···················· |
| AB | 5552 hCoV-19/England/MILK-C60F8D/2020\|EPI_ISL_769253\|20 | ···················· | ···················· | ····················· | ···················· | ······T············· | ···················· |
| AB | 5553 hCoV-19/England/QEUH-C5BBAF/2020\|EPI_ISL_769254\|20 | ···················· | ···················· | ····················· | ···················· | ···················· | ···················· |
| AB | 5554 hCoV-19/England/QEUH-C5C397/2020\|EPI_ISL_769255\|20 | ···················· | ···················· | ····················· | ···················· | ···················· | ···················· |
| AB | 5555 hCoV-19/England/MILK-C60D92/2020\|EPI_ISL_769256\|20 | ···················· | ···················· | ····················· | ···················· | ···················· | ···················· |
| AB | 5556 hCoV-19/England/MILK-C60FE7/2020\|EPI_ISL_769258\|20 | ···················· | ···················· | ····················· | ···················· | ···················· | ···················· |
| AB | 5557 hCoV-19/England/QEUH-C5BF2B/2020\|EPI_ISL_769261\|20 | ···················· | ···················· | ····················· | ···················· | ···················· | ···················· |
| AB | 5558 hCoV-19/England/QEUH-C5D563/2020\|EPI_ISL_769273\|20 | ···················· | ···················· | ····················· | ···················· | ···················· | ···················· |
| AB | 5559 hCoV-19/England/QEUH-C5D5FA/2020\|EPI_ISL_769276\|20 | ···················· | ···················· | ····················· | ···················· | ···················· | ···················· |
| AB | 5560 hCoV-19/England/MILK-C610D4/2020\|EPI_ISL_769279\|20 | ···················· | ···················· | ····················· | ···················· | ······T············· | ···················· |
| AB | 5561 hCoV-19/Canada/ON-PHL-20-03662/2020\|EPI_ISL_775100 | ···················· | ·A·················· | ····················· | ···················· | ···················· | ···A················ |
| AB | 5562 hCoV-19/Canada/ON-PHL-20-03663/2020\|EPI_ISL_775101 | ···················· | ·A·················· | ····················· | ···················· | ···················· | ···A················ |
| AB | 5563 hCoV-19/Canada/ON-PHL-20-03664/2020\|EPI_ISL_775102 | ···················· | ···················· | ····················· | ···················· | ···················· | ···················· |
| AB | 5564 hCoV-19/Canada/ON-PHL-20-03665/2020\|EPI_ISL_775103 | ···················· | ···················· | ····················· | ···················· | ···················· | ···················· |
| AB | 5565 hCoV-19/Canada/ON-PHL-20-03666/2020\|EPI_ISL_775104 | ···················· | ···················· | ····················· | ···················· | ···················· | ···················· |

| | | | | | | | | | |
|---|---|---|---|---|---|---|---|---|---|
| AB | 5567 | hCoV-19/Canada/ON-PHL-20-03747/2020\|EPI_ISL_775106 | ···················· | ···················· | ···················· | ···················· | ···················· | ···················· | |
| AB | 5568 | hCoV-19/Canada/ON-PHL-20-03748/2020\|EPI_ISL_775107 | ···················· | ···················· | ···················· | ···················· | NNN················· | ···················· | 1 |
| AB | 5569 | hCoV-19/Canada/ON-PHL-20-03749/2020\|EPI_ISL_775108 | ···················· | ···················· | ···················· | ···················· | ···················· | ···················· | |
| AB | 5570 | hCoV-19/Canada/ON-PHL-20-03750/2020\|EPI_ISL_775109 | ···················· | ·A·················· | ···················· | ···················· | ···················· | ··A················· | |
| AB | 5571 | hCoV-19/Canada/ON-PHL-20-03602/2020\|EPI_ISL_775110 | ···················· | ···················· | ···················· | ···················· | ···················· | ···················· | |
| AB | 5572 | hCoV-19/Canada/ON-PHL-20-03603/2020\|EPI_ISL_775111 | ···················· | ···················· | ···················· | ···················· | ···················· | ···················· | |
| AB | 5573 | hCoV-19/Canada/ON-PHL-20-03604/2020\|EPI_ISL_775112 | ···················· | ···················· | ···················· | ···················· | ···················· | ···················· | |
| AB | 5574 | hCoV-19/Canada/ON-PHL-20-03605/2020\|EPI_ISL_775113 | ···················· | ···················· | ···················· | ···················· | ···················· | ···················· | |
| AB | 5575 | hCoV-19/Canada/ON-PHL-20-03606/2020\|EPI_ISL_775114 | ···················· | ···················· | ···················· | ···················· | ···················· | ···················· | |
| AB | 5576 | hCoV-19/Canada/ON-PHL-20-03607/2020\|EPI_ISL_775115 | ···················· | ···················· | ···················· | ···················· | ···················· | ···················· | |
| AB | 5577 | hCoV-19/Canada/ON-PHL-20-03608/2020\|EPI_ISL_775116 | ···················· | ···················· | ···················· | ···················· | ···················· | ···················· | |
| AB | 5578 | hCoV-19/Germany/HH-hpi-p2171/2020\|EPI_ISL_776401\|2 | ···················· | ···················· | ···················· | ···················· | ···················· | ···················· | |
| AB | 5579 | hCoV-19/Germany/HH-hpi-p2174/2020\|EPI_ISL_776404\|2 | ···················· | ···················· | ···················· | ···················· | ···················· | ···················· | |
| AB | 5580 | hCoV-19/Germany/HH-hpi-p2175/2020\|EPI_ISL_776405\|2 | ···················· | ···················· | ···················· | ···················· | ···················· | ···················· | |
| AB | 5581 | hCoV-19/Germany/HH-hpi-p2177/2020\|EPI_ISL_776406\|2 | ···················· | ···················· | ···················· | ···················· | ···················· | ···················· | |
| AB | 5582 | hCoV-19/Germany/HH-hpi-p2178/2020\|EPI_ISL_776407\|2 | ···················· | ···················· | ···················· | ···················· | ···················· | ···················· | |
| AB | 5583 | hCoV-19/Germany/HH-hpi-p2179/2020\|EPI_ISL_776408\|2 | ···················· | ···················· | ···················· | ···················· | ···················· | ···················· | |
| AB | 5584 | hCoV-19/Germany/HH-hpi-p2180/2020\|EPI_ISL_776409\|2 | ···················· | ···················· | ···················· | ···················· | ···················· | ···················· | |
| AB | 5585 | hCoV-19/Germany/HH-hpi-p2181/2020\|EPI_ISL_776410\|2 | ···················· | ···················· | ···················· | ···················· | ···················· | ···················· | |
| AB | 5586 | hCoV-19/Germany/HH-hpi-p2182/2020\|EPI_ISL_776411\|2 | ···················· | ···················· | ···················· | ···················· | ···················· | ···················· | |
| AB | 5587 | hCoV-19/Germany/HH-hpi-p2183/2020\|EPI_ISL_776412\|2 | ···················· | ···················· | ···················· | ···················· | ···················· | ···················· | |
| AB | 5588 | hCoV-19/Germany/HH-hpi-p2185/2020\|EPI_ISL_776413\|2 | ···················· | ···················· | ···················· | ···················· | ···················· | ···················· | |
| AB | 5589 | hCoV-19/Germany/HH-hpi-p2186/2020\|EPI_ISL_776414\|2 | ···················· | ···················· | ···················· | ···················· | ···················· | ···················· | |
| AB | 5590 | hCoV-19/Germany/HH-hpi-p2187/2020\|EPI_ISL_776415\|2 | ···················· | ···················· | ···················· | ···················· | ···················· | ···················· | |
| AB | 5591 | hCoV-19/Germany/HH-hpi-p2188/2020\|EPI_ISL_776416\|2 | ···················· | ···················· | ···················· | ···················· | ···················· | ···················· | |
| AB | 5592 | hCoV-19/Germany/HH-hpi-p2189/2020\|EPI_ISL_776417\|2 | ···················· | ···················· | ···················· | ···················· | ···················· | ···················· | |
| AB | 5593 | hCoV-19/Germany/HH-hpi-p2190/2020\|EPI_ISL_776418\|2 | ···················· | ···················· | ···················· | ···················· | ···················· | ···················· | |
| AB | 5594 | hCoV-19/Germany/HH-hpi-p2191/2020\|EPI_ISL_776419\|2 | ···················· | ···················· | ···················· | ···················· | ···················· | ···················· | |
| AB | 5595 | hCoV-19/Germany/HH-hpi-p2192/2020\|EPI_ISL_776420\|2 | ···················· | ···················· | ···················· | ···················· | ···················· | ···················· | |
| AB | 5596 | hCoV-19/Germany/HH-hpi-p2193/2020\|EPI_ISL_776421\|2 | ···················· | ···················· | ···················· | ···················· | ···················· | ···················· | |
| AB | 5597 | hCoV-19/Germany/HH-hpi-p2194/2020\|EPI_ISL_776422\|2 | ···················· | ···················· | ···················· | ···················· | ···················· | ···················· | |
| AB | 5598 | hCoV-19/Germany/HH-hpi-p2195/2020\|EPI_ISL_776423\|2 | ···················· | ···················· | ···················· | ···················· | ···················· | ···················· | |
| AB | 5599 | hCoV-19/Germany/HH-hpi-p2196/2020\|EPI_ISL_776424\|2 | ···················· | ···················· | ···················· | ···················· | ···················· | ···················· | |
| AB | 5600 | hCoV-19/Germany/HH-hpi-p2197/2020\|EPI_ISL_776425\|2 | ···················· | ···················· | ···················· | ···················· | ···················· | ···················· | |
| AB | 5601 | hCoV-19/Germany/HH-hpi-p2198/2020\|EPI_ISL_776426\|2 | ···················· | ···················· | ···················· | ···················· | ···················· | ···················· | |
| AB | 5602 | hCoV-19/Germany/HH-hpi-p2199/2020\|EPI_ISL_776427\|2 | ···················· | ···················· | ···················· | ···················· | ···················· | ···················· | |
| AB | 5603 | hCoV-19/Germany/HH-hpi-p2295/2020\|EPI_ISL_776428\|2 | ···················· | ···················· | ···················· | ···················· | ···················· | ···················· | |
| AB | 5604 | hCoV-19/Germany/HH-hpi-p2296/2020\|EPI_ISL_776429\|2 | ···················· | ···················· | ···················· | ···················· | ···················· | ···················· | |
| AB | 5605 | hCoV-19/Germany/HH-hpi-p2297/2020\|EPI_ISL_776430\|2 | ···················· | ···················· | ···················· | ···················· | ···················· | ···················· | |
| AB | 5606 | hCoV-19/Germany/HH-hpi-p2298/2020\|EPI_ISL_776431\|2 | ···················· | ···················· | ···················· | ···················· | ···················· | ···················· | |
| AB | 5607 | hCoV-19/Germany/HH-hpi-p2299/2020\|EPI_ISL_776432\|2 | ···················· | ···················· | ···················· | ···················· | ···················· | ···················· | |
| AB | 5608 | hCoV-19/Germany/HH-hpi-p2300/2020\|EPI_ISL_776433\|2 | ···················· | ···················· | ···················· | ···················· | ···················· | ···················· | |
| AB | 5609 | hCoV-19/Germany/HH-hpi-p2302/2020\|EPI_ISL_776434\|2 | ···················· | ···················· | ···················· | ···················· | ···················· | ···················· | |
| AB | 5610 | hCoV-19/Germany/HH-hpi-p2303/2020\|EPI_ISL_776435\|2 | ···················· | ···················· | ···················· | ···················· | ···················· | ···················· | |
| AB | 5611 | hCoV-19/Germany/HH-hpi-p2304/2020\|EPI_ISL_776436\|2 | ···················· | ···················· | ···················· | ···················· | ···················· | ···················· | |
| AB | 5612 | hCoV-19/Germany/HH-hpi-p2306/2020\|EPI_ISL_776437\|2 | ···················· | ···················· | ···················· | ···················· | ···················· | ···················· | |
| AB | 5613 | hCoV-19/Germany/HH-hpi-p2307/2020\|EPI_ISL_776438\|2 | ···················· | ···················· | ···················· | ···················· | ···················· | ···················· | |
| AB | 5614 | hCoV-19/Germany/HH-hpi-p2308/2020\|EPI_ISL_776439\|2 | ···················· | ···················· | ···················· | ···················· | ···················· | ···················· | |
| AB | 5615 | hCoV-19/Germany/HH-hpi-p2309/2020\|EPI_ISL_776440\|2 | ···················· | ···················· | ···················· | ···················· | ···················· | ···················· | |
| AB | 5616 | hCoV-19/Germany/HH-hpi-p2324/2020\|EPI_ISL_776441\|2 | ···················· | ···················· | ···················· | ···················· | ···················· | ···················· | |
| AB | 5617 | hCoV-19/Germany/HH-hpi-p2325/2020\|EPI_ISL_776442\|2 | ···················· | ···A················ | ···················· | ···················· | ···················· | ···················· | |
| AB | 5618 | hCoV-19/Germany/HH-hpi-p2329/2020\|EPI_ISL_776443\|2 | ···················· | ···················· | ···················· | ···················· | ···················· | ···················· | |
| AB | 5619 | hCoV-19/Germany/HH-hpi-p2332/2020\|EPI_ISL_776444\|2 | ···················· | ···················· | ···················· | ···················· | ···················· | ···················· | |
| AB | 5620 | hCoV-19/Germany/HH-hpi-p2333/2020\|EPI_ISL_776445\|2 | ···················· | ···················· | ···················· | ···················· | ···················· | ···················· | |
| AB | 5621 | hCoV-19/Germany/HH-hpi-p2334/2020\|EPI_ISL_776446\|2 | ···················· | ···················· | ···················· | ···················· | ···················· | ···················· | |
| AB | 5622 | hCoV-19/Germany/HH-hpi-p2335/2020\|EPI_ISL_776447\|2 | ···················· | ···················· | ···················· | ···················· | ···················· | ···················· | |
| AB | 5623 | hCoV-19/England/CAMC-CB78E6/2020\|EPI_ISL_777700\|20 | ···················· | ···················· | ···················· | ···················· | ···················· | ···················· | |
| AB | 5624 | hCoV-19/England/CAMC-CB7664/2020\|EPI_ISL_777703\|20 | ···················· | ···················· | ···················· | ···················· | ···················· | ···················· | |
| AB | 5625 | hCoV-19/England/CAMC-CB6D06/2020\|EPI_ISL_777726\|20 | ···················· | ···················· | ···················· | ···················· | ···················· | ···················· | |
| AB | 5626 | hCoV-19/England/CAMC-CB8715/2020\|EPI_ISL_777728\|20 | ···················· | ···················· | ···················· | ···················· | ···················· | ···················· | |
| AB | 5627 | hCoV-19/England/CAMC-CB6D33/2020\|EPI_ISL_777737\|20 | ···················· | ···················· | ···················· | ···················· | ···················· | ···················· | |
| AB | 5628 | hCoV-19/England/CAMC-CB7E11/2020\|EPI_ISL_777743\|20 | ···················· | ···················· | ···················· | ···················· | ···················· | ···················· | |
| AB | 5629 | hCoV-19/England/CAMC-CB9F67/2020\|EPI_ISL_777744\|20 | ···················· | ···················· | ···················· | ···················· | ···················· | ···················· | |
| AB | 5630 | hCoV-19/England/CAMC-CB7EB7/2020\|EPI_ISL_777747\|20 | ···················· | ···················· | ···················· | ···················· | ···················· | ···················· | |
| AB | 5631 | hCoV-19/England/CAMC-CB8C8F/2020\|EPI_ISL_777776\|20 | ···················· | ···················· | ···················· | ···················· | ···················· | ···················· | |
| AB | 5632 | hCoV-19/England/CAMC-CB7AC2/2020\|EPI_ISL_777778\|20 | ···················· | ···················· | ···················· | ···················· | ···················· | ···················· | |
| AB | 5633 | hCoV-19/Finland/25LS7D8/2020\|EPI_ISL_759915\|2020-1 | ···················· | ·A·················· | ···················· | ···················· | ···················· | ··A················· | |
| AB | 5634 | hCoV-19/Finland/25SS15C3/2020\|EPI_ISL_759916\|2020- | ···················· | ·A·················· | ···················· | ···················· | ···················· | ··A················· | |
| AB | 5635 | hCoV-19/Finland/25SS15G11/2020\|EPI_ISL_759917\|2020 | ···················· | ·A·················· | ···················· | ···················· | ···················· | ··A················· | |
| AB | 5636 | hCoV-19/Finland/25SS22B6/2020\|EPI_ISL_759918\|2020- | ···················· | ·A·················· | ···················· | ···················· | ···················· | ··A················· | |
| AB | 5637 | hCoV-19/Finland/25SS22F2/2020\|EPI_ISL_759919\|2020- | ···················· | ···················· | ···················· | ···················· | ···················· | ···················· | |
| AB | 5638 | hCoV-19/Finland/25SS22G3/2020\|EPI_ISL_759920\|2020- | ···················· | ·A·················· | ···················· | ···················· | ···················· | ··A················· | |
| AB | 5639 | hCoV-19/Finland/25SS22H2/2020\|EPI_ISL_759921\|2020- | ···················· | ·A·················· | ···················· | ···················· | ···················· | ··A················· | |
| AB | 5640 | hCoV-19/Finland/25SS4B5/2020\|EPI_ISL_759922\|2020-0 | ···················· | ···················· | ···················· | ···················· | ···················· | ···················· | |
| AB | 5641 | hCoV-19/Finland/25SS6C12/2020\|EPI_ISL_759923\|2020- | ···················· | ·A·················· | ···················· | ···················· | ···················· | ··A················· | |
| AB | 5642 | hCoV-19/Finland/25SS6H11/2020\|EPI_ISL_759924\|2020- | ···················· | ···················· | ···················· | ···················· | ···················· | ···················· | |
| AB | 5643 | hCoV-19/Finland/25SS8D9/2020\|EPI_ISL_759925\|2020-0 | ···················· | ·A·················· | ···················· | ···················· | ···················· | ··A················· | |
| AB | 5644 | hCoV-19/Finland/25SS8E1/2020\|EPI_ISL_759926\|2020-0 | ···················· | ·A·················· | ···················· | ···················· | ···················· | ··A················· | |
| AB | 5645 | hCoV-19/Finland/25SS8H7/2020\|EPI_ISL_759927\|2020-0 | ···················· | ···················· | ···················· | ···················· | ···················· | ···················· | |
| AB | 5646 | hCoV-19/Finland/25SS9G5/2020\|EPI_ISL_759928\|2020-0 | ···················· | ·A·················· | ···················· | ···················· | ···················· | ··A················· | |

| | | | | | | | | |
|---|---|---|---|---|---|---|---|---|
| AB | 5647 | hCoV-19/Finland/26LS12CA12/2020\|EPI_ISL_759929\|202 | ···················· | ·A·················· | ···················· | ···················· | ···················· | ···A················ |
| AB | 5648 | hCoV-19/Finland/26LS12CA2/2020\|EPI_ISL_759930\|2020 | ···················· | ···················· | ···················· | ···················· | ···················· | ···················· |
| AB | 5649 | hCoV-19/Finland/26LS13CD3/2020\|EPI_ISL_759931\|2020 | ···················· | ·A·················· | ···················· | ···················· | ···················· | ···A················ |
| AB | 5650 | hCoV-19/Finland/26LS1CF9/2020\|EPI_ISL_759932\|2020- | ···················· | ·A·················· | ···················· | ···················· | ···················· | ···A················ |
| AB | 5651 | hCoV-19/Finland/26LS3CA11/2020\|EPI_ISL_759933\|2020 | ···················· | ···················· | ···················· | ···················· | ···················· | ···················· |
| AB | 5652 | hCoV-19/Finland/26LS3DB3/2020\|EPI_ISL_759934\|2020- | ···················· | ·A·················· | ···················· | ···················· | ···················· | ···A················ |
| AB | 5653 | hCoV-19/Finland/26LS3DF11/2020\|EPI_ISL_759935\|2020 | ···················· | ·A·················· | ···················· | ···················· | ···················· | ···A················ |
| AB | 5654 | hCoV-19/Finland/26LS3DF9/2020\|EPI_ISL_759936\|2020- | ···················· | ···················· | ···················· | ···················· | ···················· | ···················· |
| AB | 5655 | hCoV-19/Finland/26LS4CH5/2020\|EPI_ISL_759937\|2020- | ···················· | ·A·················· | ···················· | ···················· | ···················· | ···A················ |
| AB | 5656 | hCoV-19/Finland/26LS5CD2/2020\|EPI_ISL_759938\|2020- | ···················· | ·A·················· | ···················· | ···················· | ···················· | ···A················ |
| AB | 5657 | hCoV-19/Finland/26LS5CD4/2020\|EPI_ISL_759939\|2020- | ···················· | ·A·················· | ···················· | ···················· | ···················· | ···A················ |
| AB | 5658 | hCoV-19/Finland/26LS7CE7/2020\|EPI_ISL_759940\|2020- | ···················· | ···················· | ···················· | ···················· | ···················· | ···················· |
| AB | 5659 | hCoV-19/Finland/26LS7CF1/2020\|EPI_ISL_759941\|2020- | ···················· | ·A·················· | ···················· | ···················· | ···················· | ···A················ |
| AB | 5660 | hCoV-19/Finland/26LS7CF7/2020\|EPI_ISL_759942\|2020- | ···················· | ···················· | ···················· | ···················· | ···················· | ···················· |
| AB | 5661 | hCoV-19/Finland/26LS8CC9/2020\|EPI_ISL_759943\|2020- | ···················· | ·A·················· | ···················· | ···················· | ···················· | ···A················ |
| AB | 5662 | hCoV-19/Finland/26LS8CE9/2020\|EPI_ISL_759944\|2020- | ···················· | ···················· | ···················· | ···················· | ···················· | ···················· |
| AB | 5663 | hCoV-19/Finland/26SS12D1/2020\|EPI_ISL_759945\|2020- | ···················· | ·A·················· | ···················· | ···················· | ···················· | ···A················ |
| AB | 5664 | hCoV-19/Finland/26SS4F7/2020\|EPI_ISL_759946\|2020-0 | ···················· | ···················· | ···················· | ···················· | ···················· | ···················· |
| AB | 5665 | hCoV-19/Finland/26SS5F4/2020\|EPI_ISL_759947\|2020-0 | ···················· | ···················· | ···················· | ···················· | ···················· | ···················· |
| AB | 5666 | hCoV-19/Finland/26SS5G5/2020\|EPI_ISL_759948\|2020-0 | ···················· | ·A·················· | ···················· | ···················· | ···················· | ···A················ |
| AB | 5667 | hCoV-19/Finland/26SS6E4/2020\|EPI_ISL_759949\|2020-0 | ···················· | ·A·················· | ···················· | ···················· | ···················· | ···A················ |
| AB | 5668 | hCoV-19/Finland/26SS6H1/2020\|EPI_ISL_759950\|2020-0 | ···················· | ·A·················· | ···················· | ···················· | ···················· | ···A················ |
| AB | 5669 | hCoV-19/Finland/26SS7B1/2020\|EPI_ISL_759951\|2020-0 | ···················· | ·A·················· | ···················· | ···················· | ···················· | ···A················ |
| AB | 5670 | hCoV-19/Finland/26SS7D3/2020\|EPI_ISL_759952\|2020-0 | ···················· | ·A·················· | ···················· | ···················· | ···················· | ···A················ |
| AB | 5671 | hCoV-19/Finland/26SS9B1/2020\|EPI_ISL_759953\|2020-0 | ···················· | ·A·················· | ···················· | ···················· | ···················· | ···A················ |
| AB | 5672 | hCoV-19/Finland/26SS9F9/2020\|EPI_ISL_759954\|2020-0 | ···················· | ···················· | ···················· | ···················· | ···················· | ···················· |
| AB | 5673 | hCoV-19/Indonesia/JI-ITDua-4437NTv/2020\|EPI_ISL_75 | ···················· | ···················· | ···················· | ···················· | ···················· | ···················· |
| AB | 5674 | hCoV-19/Indonesia/JI-ITDua-5235NTv/2020\|EPI_ISL_75 | ···················· | ···················· | ···················· | ···················· | ···················· | ···················· |
| AB | 5675 | hCoV-19/Indonesia/JI-ITDua-5392NTv/2020\|EPI_ISL_75 | ···················· | ···················· | ···················· | ···················· | ···················· | ···················· |
| AB | 5676 | hCoV-19/Indonesia/JI-ITDua-5748NTv/2020\|EPI_ISL_75 | ···················· | ···················· | ···················· | ···················· | ···················· | ···················· |
| AB | 5677 | hCoV-19/England/ALDP-CB4649/2020\|EPI_ISL_760900\|20 | ···················· | ···················· | ···················· | ···················· | ···················· | ···················· |
| AB | 5678 | hCoV-19/England/CAMC-CB3158/2020\|EPI_ISL_760903\|20 | ···················· | ···················· | ···················· | ···················· | ···················· | ···················· |
| AB | 5679 | hCoV-19/England/MILK-C0B75F/2020\|EPI_ISL_760905\|20 | ···················· | ···················· | ···················· | ···················· | ···················· | ···················· |
| AB | 5680 | hCoV-19/England/ALDP-CB3932/2020\|EPI_ISL_760906\|20 | ···················· | ···················· | ···················· | ···················· | ······T············· | ···················· |
| AB | 5681 | hCoV-19/England/QEUH-CADA1A/2020\|EPI_ISL_760907\|20 | ···················· | ···················· | ···················· | ···················· | ···················· | ···················· |
| AB | 5682 | hCoV-19/England/ALDP-CB4764/2020\|EPI_ISL_760908\|20 | ···················· | ···················· | ···················· | ···················· | ···················· | ···················· |
| AB | 5683 | hCoV-19/England/QEUH-CADA56/2020\|EPI_ISL_760911\|20 | ···················· | ···················· | ···················· | ···················· | ···················· | ···················· |
| AB | 5684 | hCoV-19/England/ALDP-CB460D/2020\|EPI_ISL_760912\|20 | ···················· | ···················· | ···················· | ···················· | ······T············· | ···················· |
| AB | 5685 | hCoV-19/England/QEUH-CAD8A7/2020\|EPI_ISL_760913\|20 | ···················· | ···················· | ···················· | ···················· | ···················· | ···················· |
| AB | 5686 | hCoV-19/England/QEUH-CADAB0/2020\|EPI_ISL_760914\|20 | ···················· | ···················· | ···················· | ···················· | ···················· | ···················· |
| AB | 5687 | hCoV-19/England/ALDP-CB4290/2020\|EPI_ISL_760915\|20 | ···················· | ···················· | ···················· | ···················· | ···················· | ···················· |
| AB | 5688 | hCoV-19/England/ALDP-CB4728/2020\|EPI_ISL_760916\|20 | ···················· | ···················· | ···················· | ···················· | ···················· | ···················· |
| AB | 5689 | hCoV-19/England/ALDP-CB3CFD/2020\|EPI_ISL_760918\|20 | ···················· | ···················· | ···················· | ···················· | ······T············· | ···················· |
| AB | 5690 | hCoV-19/England/CAMC-BF9E41/2020\|EPI_ISL_760920\|20 | ···················· | ·A·················· | ···················· | ···················· | ···················· | ···················· |
| AB | 5691 | hCoV-19/England/ALDP-CB42EB/2020\|EPI_ISL_760921\|20 | ···················· | ···················· | ···················· | ···················· | ···················· | ···················· |
| AB | 5692 | hCoV-19/England/CAMC-CB2DFB/2020\|EPI_ISL_760922\|20 | ···················· | ···················· | ···················· | ···················· | ···················· | ···················· |
| AB | 5693 | hCoV-19/England/ALDP-CB3D45/2020\|EPI_ISL_760925\|20 | ···················· | ···················· | ···················· | ···················· | ···················· | ···················· |
| AB | 5694 | hCoV-19/England/QEUH-CADA65/2020\|EPI_ISL_760926\|20 | ···················· | ···················· | ···················· | ···················· | ···················· | ···················· |
| AB | 5695 | hCoV-19/England/ALDP-CB3F8B/2020\|EPI_ISL_760928\|20 | ···················· | ···················· | ···················· | ···················· | ···················· | ···················· |
| AB | 5696 | hCoV-19/England/ALDP-CB3404/2020\|EPI_ISL_760929\|20 | ···················· | ·A·················· | ···················· | ···················· | ···················· | ···················· |
| AB | 5697 | hCoV-19/England/QEUH-CADADE/2020\|EPI_ISL_760930\|20 | ···················· | ···················· | ···················· | ···················· | ···················· | ···················· |
| AB | 5698 | hCoV-19/England/CAMC-BF612D/2020\|EPI_ISL_760932\|20 | ···················· | ···················· | ···················· | ···················· | ···················· | ···················· |
| AB | 5699 | hCoV-19/England/ALDP-CB4746/2020\|EPI_ISL_760933\|20 | ···················· | ···················· | ···················· | ···················· | ···················· | ···················· |
| AB | 5700 | hCoV-19/England/QEUH-CAD1E8/2020\|EPI_ISL_760934\|20 | ···················· | ···················· | ···················· | ···················· | ···················· | ···················· |
| AB | 5701 | hCoV-19/England/QEUH-CAD0CD/2020\|EPI_ISL_760935\|20 | ···················· | ···················· | ···················· | ···················· | ···················· | ···················· |
| AB | 5702 | hCoV-19/England/CAMC-BFB146/2020\|EPI_ISL_760936\|20 | ···················· | ···················· | ···················· | ···················· | ···················· | ···················· |
| AB | 5703 | hCoV-19/England/ALDP-CB3DAF/2020\|EPI_ISL_760937\|20 | ···················· | ···················· | ···················· | ···················· | ···················· | ···················· |
| AB | 5704 | hCoV-19/England/ALDP-CB46FE/2020\|EPI_ISL_760938\|20 | ···················· | ···················· | ···················· | ···················· | ···················· | ···················· |
| AB | 5705 | hCoV-19/England/CAMC-CB256C/2020\|EPI_ISL_760939\|20 | ···················· | ···················· | ···················· | ···················· | ···················· | ···················· |
| AB | 5706 | hCoV-19/Italy/UMB-IZSUM-48682-7/2020\|EPI_ISL_76313 | ···················· | ···················· | ···················· | ···················· | ···················· | ···················· |
| AB | 5707 | hCoV-19/Netherlands/NB-EMC-481/2020\|EPI_ISL_763140 | ···················· | ···················· | ···················· | ···················· | ···················· | ···················· |
| AB | 5708 | hCoV-19/Netherlands/NB-EMC-482/2020\|EPI_ISL_763141 | ···················· | ···················· | ···················· | ···················· | ···················· | ···················· |
| AB | 5709 | hCoV-19/Netherlands/NB-EMC-483/2020\|EPI_ISL_763142 | ···················· | ···················· | ···················· | ···················· | ···················· | ···················· |
| AB | 5710 | hCoV-19/Netherlands/NB-EMC-484/2020\|EPI_ISL_763143 | ···················· | ···················· | ···················· | ···················· | ···················· | ···················· |
| AB | 5711 | hCoV-19/Netherlands/NB-EMC-485/2020\|EPI_ISL_763144 | ···················· | ···················· | ···················· | ···················· | ···················· | ···················· |
| AB | 5712 | hCoV-19/Netherlands/NB-EMC-486/2020\|EPI_ISL_763145 | ···················· | ···················· | ···················· | ···················· | ···················· | ···················· |
| AB | 5713 | hCoV-19/Netherlands/NB-EMC-488/2020\|EPI_ISL_763147 | ···················· | ···················· | ···················· | ···················· | ···················· | ···················· |
| AB | 5714 | hCoV-19/Netherlands/NB-EMC-489/2020\|EPI_ISL_763148 | ···················· | ···················· | ···················· | ···················· | ···················· | ···················· |
| AB | 5715 | hCoV-19/Netherlands/NB-EMC-490/2020\|EPI_ISL_763149 | ···················· | ···················· | ···················· | ···················· | ···················· | ···················· |
| AB | 5716 | hCoV-19/Netherlands/OV-EMC-110/2020\|EPI_ISL_763150 | ···················· | ···················· | ···················· | ···················· | ···················· | ···················· |
| AB | 5717 | hCoV-19/Netherlands/OV-EMC-111/2020\|EPI_ISL_763151 | ···················· | ······A············· | ···················· | ···················· | ···················· | ···················· |
| AB | 5718 | hCoV-19/Netherlands/OV-EMC-112/2020\|EPI_ISL_763152 | ···················· | ···················· | ···················· | ···················· | ···················· | ···················· |
| AB | 5719 | hCoV-19/Netherlands/OV-EMC-113/2020\|EPI_ISL_763153 | ···················· | ···················· | ···················· | ···················· | ···················· | ···················· |
| AB | 5720 | hCoV-19/Netherlands/OV-EMC-115/2020\|EPI_ISL_763155 | ···················· | ···················· | ···················· | ···················· | ···················· | ···················· |
| AB | 5721 | hCoV-19/Netherlands/OV-EMC-116/2020\|EPI_ISL_763156 | ···················· | ···················· | ···················· | ···················· | ···················· | ···················· |
| AB | 5722 | hCoV-19/Netherlands/OV-EMC-117/2020\|EPI_ISL_763157 | ···················· | ···················· | ···················· | ···················· | ···················· | ···················· |
| AB | 5723 | hCoV-19/Netherlands/OV-EMC-118/2020\|EPI_ISL_763158 | ···················· | ···················· | ···················· | ···················· | ···················· | ···················· |
| AB | 5724 | hCoV-19/Netherlands/OV-EMC-119/2020\|EPI_ISL_763159 | ···T················ | ···················· | ···················· | ···················· | ···················· | ···················· |
| AB | 5725 | hCoV-19/Netherlands/OV-EMC-120/2020\|EPI_ISL_763160 | ···················· | ···················· | ···················· | ···················· | ···················· | ···················· |
| AB | 5726 | hCoV-19/Netherlands/OV-EMC-122/2020\|EPI_ISL_763162 | ···················· | ···················· | ···················· | ···················· | ···················· | ···················· |

| | | | | | | | |
|---|---|---|---|---|---|---|---|
| AB | 5727 hCoV-19/Netherlands/un-EMC-827/2020\|EPI_ISL_763180 | ···················· | ···················· | ···················· | ···················· | ···················· | ···················· |
| AB | 5728 hCoV-19/Netherlands/UT-EMC-230/2020\|EPI_ISL_763182 | ···················· | ···················· | ···················· | ···················· | ···················· | ···················· |
| AB | 5729 hCoV-19/Netherlands/UT-EMC-231/2020\|EPI_ISL_763183 | ···················· | ···················· | ···················· | ···················· | ···················· | ···················· |
| AB | 5730 hCoV-19/Netherlands/UT-EMC-232/2020\|EPI_ISL_763184 | ···················· | ···················· | ···················· | ···················· | ···················· | ···················· |
| AB | 5731 hCoV-19/Netherlands/UT-EMC-233/2020\|EPI_ISL_763185 | ···················· | ···················· | ···················· | ···················· | ···················· | ···················· |
| AB | 5732 hCoV-19/Netherlands/UT-EMC-234/2020\|EPI_ISL_763186 | ···················· | ···················· | ···················· | ···················· | ···················· | ···················· |
| AB | 5733 hCoV-19/Netherlands/UT-EMC-235/2020\|EPI_ISL_763187 | ···················· | ···················· | ···················· | ···················· | ···················· | ···················· |
| AB | 5734 hCoV-19/Netherlands/ZE-EMC-88/2020\|EPI_ISL_763189\| | ···················· | ···················· | ···················· | ···················· | ···················· | ···················· |
| AB | 5735 hCoV-19/Netherlands/ZE-EMC-89/2020\|EPI_ISL_763190\| | ···················· | ···················· | ···················· | ···················· | ···················· | ···················· |
| AB | 5736 hCoV-19/Netherlands/ZH-EMC-1112/2020\|EPI_ISL_76319 | ···················· | ···················· | ···················· | ···················· | ···················· | ···················· |
| AB | 5737 hCoV-19/Netherlands/ZH-EMC-1113/2020\|EPI_ISL_76319 | ···················· | ···················· | ···················· | ···················· | ···················· | ···················· |
| AB | 5738 hCoV-19/Netherlands/ZH-EMC-1114/2020\|EPI_ISL_76319 | ···················· | ···················· | ···················· | ···················· | ···················· | ···················· |
| AB | 5739 hCoV-19/Netherlands/ZH-EMC-1115/2020\|EPI_ISL_76319 | ···················· | ···················· | ···················· | ···················· | ···················· | ···················· |
| AB | 5740 hCoV-19/Netherlands/ZH-EMC-1119/2020\|EPI_ISL_76319 | ···················· | ···················· | ···················· | ···················· | ···················· | ···················· |
| AB | 5741 hCoV-19/Netherlands/ZH-EMC-1120/2020\|EPI_ISL_76319 | ···················· | ···················· | ···················· | ···················· | ···················· | ···················· |
| AB | 5742 hCoV-19/Northern Ireland/NIRE-23512B/2020\|EPI_ISL_ | ···················· | ···················· | ···················· | ···················· | ···················· | ···················· |
| AB | 5744 hCoV-19/Northern Ireland/NIRE-2351C1/2020\|EPI_ISL_ | ···················· | ···················· | ···················· | ···················· | ···················· | ···················· |
| AB | 5745 hCoV-19/Northern Ireland/NIRE-2351FE/2020\|EPI_ISL_ | ···················· | ···················· | ···················· | ···················· | ···················· | ···················· |
| AB | 5746 hCoV-19/Northern Ireland/NIRE-23520A/2020\|EPI_ISL_ | ···················· | ···················· | ···················· | ···················· | ···················· | ···················· |
| AB | 5748 hCoV-19/Northern Ireland/NIRE-235228/2020\|EPI_ISL_ | ···················· | ···················· | ···················· | ···················· | ···················· | ···················· |
| AB | 5749 hCoV-19/Northern Ireland/NIRE-235237/2020\|EPI_ISL_ | ···················· | ···················· | ···················· | ···················· | ···················· | ···················· |
| AB | 5751 hCoV-19/Northern Ireland/NIRE-235273/2020\|EPI_ISL_ | ···················· | ···················· | ···················· | ···················· | ···················· | ···················· |
| AB | 5753 hCoV-19/England/NORW-221C03/2020\|EPI_ISL_764469\|20 | ···················· | ···················· | ···················· | ···················· | ···················· | ···················· |
| AB | 5754 hCoV-19/England/NORW-221C12/2020\|EPI_ISL_764470\|20 | ···················· | ···················· | ···················· | ···················· | ···················· | ···················· |
| AB | 5755 hCoV-19/England/NORW-221C4F/2020\|EPI_ISL_764471\|20 | ···················· | ···················· | ···················· | ···················· | ···················· | ···················· |
| AB | 5756 hCoV-19/England/NORW-221C5E/2020\|EPI_ISL_764472\|20 | ···················· | ···················· | ···················· | ···················· | ···················· | ···················· |
| AB | 5757 hCoV-19/England/NORW-221C7C/2020\|EPI_ISL_764473\|20 | ···················· | ···················· | ···················· | ···················· | ···················· | ···················· |
| AB | 5758 hCoV-19/England/NORW-221D5B/2020\|EPI_ISL_764474\|20 | ···················· | ···················· | ···················· | ···················· | ···················· | ···················· |
| AB | 5759 hCoV-19/England/NORW-221DD3/2020\|EPI_ISL_764475\|20 | ···················· | ···················· | ···················· | ···················· | ···················· | ···················· |
| AB | 5760 hCoV-19/England/NORW-221DE2/2020\|EPI_ISL_764476\|20 | ···················· | ···················· | ···················· | ···················· | ···················· | ···················· |
| AB | 5761 hCoV-19/England/NORW-221E76/2020\|EPI_ISL_764477\|20 | ···················· | ···················· | ···················· | ···················· | ···················· | ···················· |
| AB | 5762 hCoV-19/England/NORW-221E94/2020\|EPI_ISL_764478\|20 | ···················· | ···················· | ···················· | ···················· | ···················· | ···················· |
| AB | 5763 hCoV-19/England/NORW-221EC1/2020\|EPI_ISL_764479\|20 | ···················· | ···················· | ···················· | ···················· | ···················· | ···················· |
| AB | 5764 hCoV-19/England/NORW-221ED0/2020\|EPI_ISL_764480\|20 | ···················· | ···················· | ···················· | ···················· | ···················· | ···················· |
| AB | 5765 hCoV-19/England/NORW-221EEF/2020\|EPI_ISL_764481\|20 | ···················· | ···················· | ···················· | ···················· | ···················· | ···················· |
| AB | 5766 hCoV-19/England/NORW-221F55/2020\|EPI_ISL_764482\|20 | ···················· | ···················· | ···················· | ···················· | ···················· | ···················· |
| AB | 5767 hCoV-19/England/NORW-221F64/2020\|EPI_ISL_764483\|20 | ···················· | ···················· | ···················· | ···················· | ···················· | ···················· |
| AB | 5768 hCoV-19/England/NORW-221F82/2020\|EPI_ISL_764484\|20 | ···················· | ···················· | ···················· | ···················· | ···················· | ···················· |
| AB | 5769 hCoV-19/England/NORW-2222E2/2020\|EPI_ISL_764485\|20 | ···················· | ···················· | ···················· | ···················· | ···················· | ···················· |
| AB | 5770 hCoV-19/England/NORW-22232B/2020\|EPI_ISL_764486\|20 | ···················· | ···················· | ···················· | ···················· | ···················· | ···················· |
| AB | 5771 hCoV-19/England/NORW-22233A/2020\|EPI_ISL_764487\|20 | ···················· | ···················· | ···················· | ···················· | ···················· | ···················· |
| AB | 5772 hCoV-19/England/NORW-2223FE/2020\|EPI_ISL_764488\|20 | ···················· | ···················· | ···················· | ···················· | ···················· | ···················· |
| AB | 5773 hCoV-19/England/NORW-222446/2020\|EPI_ISL_764489\|20 | ···················· | ···················· | ···················· | ···················· | ···················· | ···················· |
| AB | 5774 hCoV-19/England/NORW-222525/2020\|EPI_ISL_764490\|20 | ···················· | ···················· | ···················· | ···················· | ···················· | ···················· |
| AB | 5775 hCoV-19/England/NORW-222543/2020\|EPI_ISL_764491\|20 | ···················· | ···················· | ···················· | ···················· | ···················· | ···················· |
| AB | 5776 hCoV-19/England/NORW-222895/2020\|EPI_ISL_764492\|20 | ···················· | ···················· | ···················· | ···················· | ···················· | ···················· |
| AB | 5777 hCoV-19/England/NOTT-121786/2020\|EPI_ISL_764493\|20 | ···················· | ···················· | ···················· | ···················· | ···················· | ···················· |
| AB | 5778 hCoV-19/USA/MA-MGH-03203/2020\|EPI_ISL_765745\|2020- | ···················· | ···················· | ···················· | ···················· | ···················· | ···················· |
| AB | 5779 hCoV-19/USA/MA-MGH-03205/2020\|EPI_ISL_765746\|2020- | ···················· | ···················· | ···················· | ···················· | ···················· | ···················· |
| AB | 5780 hCoV-19/USA/MA-MGH-03212/2020\|EPI_ISL_765747\|2020- | ···················· | ···················· | ···················· | ···················· | ···················· | ···················· |
| AB | 5781 hCoV-19/USA/MA-MGH-03216/2020\|EPI_ISL_765748\|2020- | ···················· | ···················· | ···················· | ···················· | ···················· | ···················· |
| AB | 5782 hCoV-19/USA/MA-MGH-03217/2020\|EPI_ISL_765749\|2020- | ···················· | ···················· | ···················· | ···················· | ···················· | ···················· |
| AB | 5783 hCoV-19/USA/MA-MGH-03220/2020\|EPI_ISL_765750\|2020- | ···················· | ···················· | ···················· | ···················· | ···················· | ···················· |
| AB | 5784 hCoV-19/USA/MA-MGH-03230/2020\|EPI_ISL_765751\|2020- | ···················· | ···················· | ···················· | ···················· | ···················· | ···················· |
| AB | 5785 hCoV-19/USA/MA-MGH-03232/2020\|EPI_ISL_765752\|2020- | ···················· | ···················· | ···················· | ···················· | ···················· | ···················· |
| AB | 5786 hCoV-19/USA/MA-MGH-03239/2020\|EPI_ISL_765754\|2020- | ···················· | ···················· | ···················· | ···················· | ···················· | ···················· |
| AB | 5787 hCoV-19/USA/MA-MGH-03242/2020\|EPI_ISL_765755\|2020- | ···················· | ·A·················· | ···················· | ···················· | ···················· | ···················· |
| AB | 5788 hCoV-19/Canada/ON-PHL-20-03609/2020\|EPI_ISL_775117 | ···················· | ···················· | ···················· | ···················· | ···················· | ···················· |
| AB | 5789 hCoV-19/Canada/ON-PHL-20-03610/2020\|EPI_ISL_775118 | ···················· | ···················· | ···················· | ···················· | ···················· | ···················· |
| AB | 5791 hCoV-19/Canada/ON-PHL-20-03612/2020\|EPI_ISL_775120 | ···················· | ···················· | ···················· | ···················· | ···················· | ···················· |
| AB | 5792 hCoV-19/Canada/ON-PHL-20-03613/2020\|EPI_ISL_775121 | ···················· | ···················· | ···················· | ···················· | ···················· | ···················· |
| AB | 5793 hCoV-19/Canada/ON-PHL-20-03615/2020\|EPI_ISL_775122 | ···················· | ···················· | ···················· | ···················· | ···················· | ···················· |
| AB | 5794 hCoV-19/Canada/ON-PHL-20-03616/2020\|EPI_ISL_775123 | ···················· | ···················· | ···················· | ···················· | ···················· | ···················· |
| AB | 5795 hCoV-19/Canada/ON-PHL-20-03617/2020\|EPI_ISL_775124 | ···················· | ···················· | ···················· | ···················· | ···················· | ···················· |
| AB | 5796 hCoV-19/Canada/ON-PHL-20-03618/2020\|EPI_ISL_775125 | ···················· | ·A·················· | ···················· | ···················· | ···················· | ··A················· |
| AB | 5797 hCoV-19/Canada/ON-PHL-20-03619/2020\|EPI_ISL_775126 | ···················· | ·A·················· | ···················· | ···················· | ···················· | ··A················· |
| AB | 5798 hCoV-19/Canada/ON-PHL-20-03621/2020\|EPI_ISL_775128 | ···················· | ···················· | ···················· | ···················· | ···················· | ···················· |
| AB | 5799 hCoV-19/Canada/ON-PHL-20-03622/2020\|EPI_ISL_775129 | ···················· | ·A·················· | ···················· | ···················· | ···················· | ··A················· |
| AB | 5800 hCoV-19/Canada/ON-PHL-20-03623/2020\|EPI_ISL_775130 | ···················· | ···················· | ···················· | ···················· | ···················· | ···················· |
| AB | 5801 hCoV-19/Canada/ON-PHL-20-03625/2020\|EPI_ISL_775131 | ···················· | ···················· | ···················· | ···················· | ···················· | ···················· |
| AB | 5802 hCoV-19/Canada/ON-PHL-20-03626/2020\|EPI_ISL_775132 | ···················· | ···················· | ···················· | ···················· | ···················· | ···················· |
| AB | 5803 hCoV-19/Canada/ON-PHL-20-03627/2020\|EPI_ISL_775133 | ···················· | ···················· | ···················· | ···················· | ···················· | ···················· |
| AB | 5804 hCoV-19/Canada/ON-PHL-20-03628/2020\|EPI_ISL_775134 | ···················· | ···················· | ···················· | ···················· | ···················· | ···················· |
| AB | 5805 hCoV-19/Canada/ON-PHL-20-03629/2020\|EPI_ISL_775135 | ···················· | ·A·················· | ···················· | ···················· | ···················· | ··A················· |
| AB | 5806 hCoV-19/Canada/ON-PHL-20-03633/2020\|EPI_ISL_775137 | ···················· | ···················· | ···················· | ···················· | ···················· | ···················· |
| AB | 5807 hCoV-19/Canada/ON-PHL-20-03634/2020\|EPI_ISL_775138 | ···················· | ·A·················· | ···················· | ···················· | ···················· | ··A················· |
| AB | 5808 hCoV-19/Canada/ON-PHL-20-03676/2020\|EPI_ISL_775160 | ···················· | ·A·················· | ···················· | ···················· | ···················· | ··A················· |
| AB | 5809 hCoV-19/Canada/ON-PHL-20-03677/2020\|EPI_ISL_775161 | ···················· | ···················· | ···················· | ···················· | ···················· | ···················· |
| AB | 5811 hCoV-19/Canada/ON-PHL-20-03679/2020\|EPI_ISL_775163 | ···················· | ···················· | ···················· | ···················· | ···················· | ···················· |
| AB | 5812 hCoV-19/Canada/ON-PHL-20-03680/2020\|EPI_ISL_775164 | ···················· | ···················· | ···················· | ···················· | ···················· | ···················· |

| | | | | | | | | |
|---|---|---|---|---|---|---|---|---|
| AB | 5813 | hCoV-19/Canada/ON-PHL-20-03681/2020\|EPI_ISL_775165 | ···················· | ···················· | ···················· | ···················· | ···················· | ···················· |
| AB | 5814 | hCoV-19/Canada/ON-PHL-20-03683/2020\|EPI_ISL_775166 | ···················· | ···················· | ···················· | ···················· | ···················· | ···················· |
| AB | 5815 | hCoV-19/Canada/ON-PHL-20-03684/2020\|EPI_ISL_775167 | ···················· | ···················· | ···················· | ···················· | ···················· | ···················· |
| AB | 5816 | hCoV-19/Canada/ON-PHL-20-03685/2020\|EPI_ISL_775168 | ···················· | ·A·················· | ···················· | ···················· | ···················· | ··A················· |
| AB | 5817 | hCoV-19/Canada/ON-PHL-20-03686/2020\|EPI_ISL_775169 | ···················· | ·A·················· | ···················· | ···················· | ···················· | ··A················· |
| AB | 5818 | hCoV-19/Canada/ON-PHL-20-03687/2020\|EPI_ISL_775170 | ···················· | ·A·················· | ···················· | ···················· | ···················· | ··A················· |
| AB | 5819 | hCoV-19/Canada/ON-PHL-20-03688/2020\|EPI_ISL_775171 | ···················· | ·A·················· | ···················· | ···················· | ···················· | ··A················· |
| AB | 5820 | hCoV-19/Canada/ON-PHL-20-03689/2020\|EPI_ISL_775172 | ···················· | ·A·················· | ···················· | ···················· | ···················· | ··A················· |
| AB | 5821 | hCoV-19/Canada/ON-PHL-20-03690/2020\|EPI_ISL_775173 | ···················· | ·A·················· | ···················· | ···················· | ···················· | ··A················· |
| AB | 5822 | hCoV-19/Canada/ON-PHL-20-03691/2020\|EPI_ISL_775174 | ···················· | ···················· | ···················· | ···················· | ···················· | ···················· |
| AB | 5823 | hCoV-19/Canada/ON-PHL-20-03692/2020\|EPI_ISL_775175 | ···················· | ···················· | ···················· | ···················· | ···················· | ···················· |
| AB | 5824 | hCoV-19/Canada/ON-PHL-20-03693/2020\|EPI_ISL_775176 | ···················· | ···················· | ···················· | ···················· | ···················· | ···················· |
| AB | 5825 | hCoV-19/Canada/ON-PHL-20-03694/2020\|EPI_ISL_775177 | ···················· | ···················· | ···················· | ···················· | ···················· | ···················· |
| AB | 5826 | hCoV-19/Canada/ON-PHL-20-03695/2020\|EPI_ISL_775178 | ···················· | ···················· | ···················· | ···················· | ···················· | ···················· |
| AB | 5827 | hCoV-19/Canada/ON-PHL-20-03696/2020\|EPI_ISL_775179 | ···················· | ···················· | ···················· | ···················· | ···················· | ···················· |
| AB | 5828 | hCoV-19/Canada/ON-PHL-20-03697/2020\|EPI_ISL_775180 | ···················· | ···················· | ···················· | ···················· | ···················· | ···················· |
| AB | 5829 | hCoV-19/Canada/ON-PHL-20-03698/2020\|EPI_ISL_775181 | ···················· | ···················· | ···················· | ···················· | ···················· | ···················· |
| AB | 5830 | hCoV-19/Canada/ON-PHL-20-03699/2020\|EPI_ISL_775182 | ···················· | ···················· | ···················· | ···················· | ···················· | ···················· |
| AB | 5831 | hCoV-19/Canada/ON-PHL-20-03700/2020\|EPI_ISL_775183 | ···················· | ···················· | ···················· | ···················· | ···················· | ···················· |
| AB | 5832 | hCoV-19/Canada/ON-PHL-20-03701/2020\|EPI_ISL_775184 | ···················· | ···················· | ···················· | ···················· | ···················· | ···················· |
| AB | 5833 | hCoV-19/Canada/ON-PHL-20-03702/2020\|EPI_ISL_775185 | ···················· | ···················· | ···················· | ···················· | ···················· | ···················· |
| AB | 5834 | hCoV-19/Canada/ON-PHL-20-03703/2020\|EPI_ISL_775186 | ···················· | ···················· | ···················· | ···················· | ···················· | ···················· |
| AB | 5835 | hCoV-19/Canada/ON-PHL-20-03704/2020\|EPI_ISL_775187 | ···················· | ·A·················· | ···················· | ···················· | ···················· | ··A················· |
| AB | 5836 | hCoV-19/Canada/ON-PHL-20-03705/2020\|EPI_ISL_775188 | ···················· | ···················· | ···················· | ···················· | ···················· | ···················· |
| AB | 5838 | hCoV-19/Canada/ON-PHL-20-03708/2020\|EPI_ISL_775190 | ···················· | ·A·················· | ···················· | ···················· | ···················· | ··A················· |
| AB | 5839 | hCoV-19/Canada/ON-PHL-20-03709/2020\|EPI_ISL_775191 | ···················· | ···················· | ···················· | ···················· | ···················· | ···················· |
| AB | 5840 | hCoV-19/Canada/ON-PHL-20-03710/2020\|EPI_ISL_775192 | ···················· | ···················· | ···················· | ···················· | ···················· | ···················· |
| AB | 5841 | hCoV-19/Canada/ON-PHL-20-03711/2020\|EPI_ISL_775193 | ···················· | ···················· | ···················· | ···················· | ···················· | ···················· |
| AB | 5842 | hCoV-19/Canada/ON-PHL-20-03712/2020\|EPI_ISL_775194 | ···················· | ···················· | ···················· | ···················· | ···················· | ···················· |
| AB | 5843 | hCoV-19/Canada/ON-PHL-20-03713/2020\|EPI_ISL_775195 | ···················· | ·A·················· | ···················· | ···················· | ···················· | ··A················· |
| AB | 5845 | hCoV-19/Canada/ON-PHL-20-03715/2020\|EPI_ISL_775197 | ···················· | ···················· | ···················· | ···················· | ···················· | ···················· |
| AB | 5846 | hCoV-19/Canada/ON-PHL-20-03716/2020\|EPI_ISL_775198 | ···················· | ·A·················· | ···················· | ···················· | ···················· | ··A················· |
| AB | 5847 | hCoV-19/Germany/HH-hpi-p2170/2020\|EPI_ISL_776400\|2 | ···················· | ···················· | ···················· | ···················· | ···················· | ···················· |
| AB | 5848 | hCoV-19/Germany/HH-hpi-p2172/2020\|EPI_ISL_776402\|2 | ···················· | ···················· | ···················· | ···················· | ···················· | ···················· |
| AB | 5849 | hCoV-19/Germany/HH-hpi-p2173/2020\|EPI_ISL_776403\|2 | ···················· | ···················· | ···················· | ···················· | ···················· | ···················· |
| AB | 5850 | hCoV-19/Germany/HH-hpi-p2338/2020\|EPI_ISL_776448\|2 | ···················· | ···················· | ···················· | ···················· | ···················· | ···················· |
| AB | 5852 | hCoV-19/Germany/HH-hpi-p2359/2020\|EPI_ISL_776450\|2 | ···················· | ···················· | ···················· | ···················· | ···················· | ···················· |
| AB | 5853 | hCoV-19/Germany/HH-hpi-p2361/2020\|EPI_ISL_776451\|2 | ···················· | ···················· | ···················· | ···················· | ···················· | ···················· |
| AB | 5854 | hCoV-19/Germany/HH-hpi-p2364/2020\|EPI_ISL_776452\|2 | ···················· | ···················· | ···················· | ···················· | ···················· | ···················· |
| AB | 5855 | hCoV-19/Germany/HH-hpi-p2365/2020\|EPI_ISL_776453\|2 | ···················· | ···················· | ···················· | ···················· | ···················· | ···················· |
| AB | 5856 | hCoV-19/Germany/HH-hpi-p2367/2020\|EPI_ISL_776454\|2 | ···················· | ···················· | ···················· | ···················· | ···················· | ···················· |
| AB | 5857 | hCoV-19/Germany/HH-hpi-p2376/2020\|EPI_ISL_776455\|2 | ···················· | ···················· | ···················· | ···················· | ···················· | ···················· |
| AB | 5858 | hCoV-19/Germany/HH-hpi-p2378/2020\|EPI_ISL_776456\|2 | ···················· | ···················· | ···················· | ···················· | ···················· | ···················· |
| AB | 5859 | hCoV-19/Germany/HH-hpi-p2379/2020\|EPI_ISL_776457\|2 | ···················· | ···················· | ···················· | ···················· | ···················· | ···················· |
| AB | 5860 | hCoV-19/Germany/HH-hpi-p2380/2020\|EPI_ISL_776458\|2 | ···················· | ···················· | ···················· | ···················· | ···················· | ···················· |
| AB | 5861 | hCoV-19/Germany/HH-hpi-p2381/2020\|EPI_ISL_776459\|2 | ···················· | ···················· | ···················· | ···················· | ···················· | ···················· |
| AB | 5862 | hCoV-19/Germany/HH-hpi-p2382/2020\|EPI_ISL_776460\|2 | ···················· | ···················· | ···················· | ···················· | ···················· | ···················· |
| AB | 5864 | hCoV-19/Germany/HH-hpi-p2384/2020\|EPI_ISL_776462\|2 | ···················· | ···················· | ···················· | ···················· | ···················· | ···················· |
| AB | 5865 | hCoV-19/Germany/HH-hpi-p2385/2020\|EPI_ISL_776463\|2 | ···················· | ···················· | ···················· | ···················· | ···················· | ···················· |
| AB | 5866 | hCoV-19/Germany/HH-hpi-p2386/2020\|EPI_ISL_776464\|2 | ···················· | ···················· | ···················· | ···················· | ···················· | ···················· |
| AB | 5868 | hCoV-19/Germany/HH-hpi-p2390/2020\|EPI_ISL_776466\|2 | ···················· | ···················· | ···················· | ···················· | ···················· | ···················· |
| AB | 5869 | hCoV-19/Germany/HH-hpi-p2392/2020\|EPI_ISL_776467\|2 | ···················· | ···················· | ···················· | ···················· | ···················· | ···················· |
| AB | 5870 | hCoV-19/Germany/HH-hpi-p2393/2020\|EPI_ISL_776468\|2 | ···················· | ···················· | ···················· | ···················· | ···················· | ···················· |
| AB | 5871 | hCoV-19/Germany/HH-hpi-p2394/2020\|EPI_ISL_776469\|2 | ···················· | ···················· | ···················· | ···················· | ···················· | ···················· |
| AB | 5872 | hCoV-19/Germany/HH-hpi-p2421/2020\|EPI_ISL_776490\|2 | ···················· | ···················· | ···················· | ···················· | ···················· | ···················· |
| AB | 5873 | hCoV-19/Germany/HH-hpi-p2422/2020\|EPI_ISL_776491\|2 | ···················· | ···················· | ···················· | ···················· | ···················· | ···················· |
| AB | 5874 | hCoV-19/Germany/HH-hpi-p2423/2020\|EPI_ISL_776492\|2 | ···················· | ···················· | ···················· | ···················· | ···················· | ···················· |
| AB | 5875 | hCoV-19/Germany/HH-hpi-p2424/2020\|EPI_ISL_776493\|2 | ···················· | ···················· | ···················· | ···················· | ···················· | ···················· |
| AB | 5876 | hCoV-19/Germany/HH-hpi-p2425/2020\|EPI_ISL_776494\|2 | ···················· | ···················· | ···················· | ···················· | ···················· | ···················· |
| AB | 5877 | hCoV-19/Germany/HH-hpi-p2426/2020\|EPI_ISL_776495\|2 | ···················· | ···················· | ···················· | ···················· | ···················· | ···················· |
| AB | 5878 | hCoV-19/Germany/HH-hpi-p2427/2020\|EPI_ISL_776496\|2 | ···················· | ···················· | ···················· | ···················· | ···················· | ···················· |
| AB | 5879 | hCoV-19/Germany/HH-hpi-p2428/2020\|EPI_ISL_776497\|2 | ···················· | ···················· | ···················· | ···················· | ···················· | ···················· |
| AB | 5880 | hCoV-19/Germany/HH-hpi-p2429/2020\|EPI_ISL_776498\|2 | ···················· | ···················· | ···················· | ···················· | ···················· | ···················· |
| AB | 5881 | hCoV-19/Germany/HH-hpi-p2430/2020\|EPI_ISL_776499\|2 | ···················· | ···················· | ···················· | ···················· | ···················· | ···················· |
| AB | 5882 | hCoV-19/England/CAMC-CBA19D/2020\|EPI_ISL_777715\|20 | ···················· | ·A·················· | ···················· | ···················· | ···················· | ···················· |
| AB | 5883 | hCoV-19/England/CAMC-CB70FD/2020\|EPI_ISL_777718\|20 | ···················· | ···················· | ···················· | ···················· | ···················· | ···················· |
| AB | 5884 | hCoV-19/England/CAMC-CBA388/2020\|EPI_ISL_777779\|20 | ···················· | ···A················ | ···················· | ···················· | ···················· | ···················· |
| AB | 5885 | hCoV-19/England/CAMC-CB7A95/2020\|EPI_ISL_777780\|20 | ···················· | ···················· | ···················· | ···················· | ···················· | ···················· |
| AB | 5886 | hCoV-19/England/CAMC-CB87F7/2020\|EPI_ISL_777781\|20 | ···················· | ···················· | ···················· | ···················· | ···················· | ···················· |
| AB | 5887 | hCoV-19/England/CAMC-CB7558/2020\|EPI_ISL_777789\|20 | ···················· | ···················· | ···················· | ···················· | ·····T·············· | ···················· |
| AB | 5888 | hCoV-19/England/CAMC-CB79B6/2020\|EPI_ISL_777796\|20 | ···················· | ···················· | ···················· | ···················· | ···················· | ···················· |
| AB | 5889 | hCoV-19/Finland/25LS10CB3/2020\|EPI_ISL_759900\|2020 | ···················· | ·A·················· | ···················· | ···················· | ···················· | ··A················· |
| AB | 5890 | hCoV-19/Finland/25LS10CE11/2020\|EPI_ISL_759901\|202 | ···················· | ·A·················· | ···················· | ···················· | ···················· | ··A················· |
| AB | 5891 | hCoV-19/Finland/25LS10CF5/2020\|EPI_ISL_759902\|2020 | ···················· | ···················· | ···················· | ···················· | ···················· | ···················· |
| AB | 5892 | hCoV-19/Finland/25LS11CE1/2020\|EPI_ISL_759903\|2020 | ···················· | ·A·················· | ···················· | ···················· | ···················· | ··A················· |
| AB | 5893 | hCoV-19/Finland/25LS11CF5/2020\|EPI_ISL_759904\|2020 | ···················· | ···················· | ···················· | ···················· | ···················· | ···················· |
| AB | 5894 | hCoV-19/Finland/25LS11CH3/2020\|EPI_ISL_759905\|2020 | ···················· | ·A·················· | ···················· | ···················· | ···················· | ··A················· |
| AB | 5895 | hCoV-19/Finland/25LS13D11/2020\|EPI_ISL_759906\|2020 | ···················· | ···················· | ···················· | ···················· | ···················· | ···················· |
| AB | 5896 | hCoV-19/Finland/25LS13D5/2020\|EPI_ISL_759907\|2020- | ···················· | ···················· | ···················· | ···················· | ···················· | ···················· |
| AB | 5897 | hCoV-19/Finland/25LS13G5/2020\|EPI_ISL_759908\|2020- | ···················· | ·A·················· | ···················· | ···················· | ···················· | ··A················· |

| | | | | | | | | |
|---|---|---|---|---|---|---|---|---|
| AB | 5898 | hCoV-19/Finland/25LS13H1/2020\|EPI_ISL_759909\|2020- | ···················· | ·A·················· | ····················· | ···················· | ···················· | ··A················· |
| AB | 5899 | hCoV-19/Finland/25LS1DB5/2020\|EPI_ISL_759910\|2020- | ···················· | ·A·················· | ····················· | ···················· | ···················· | ··A················· |
| AB | 5900 | hCoV-19/Finland/25LS1DC1/2020\|EPI_ISL_759911\|2020- | ···················· | ·A·················· | ····················· | ···················· | ···················· | ··A················· |
| AB | 5901 | hCoV-19/Finland/25LS6CB7/2020\|EPI_ISL_759912\|2020- | ···················· | ···················· | ····················· | ···················· | ···················· | ···················· |
| AB | 5902 | hCoV-19/Finland/25LS6CE1/2020\|EPI_ISL_759913\|2020- | ···················· | ···················· | ····················· | ···················· | ···················· | ···················· |
| AB | 5903 | hCoV-19/Finland/25LS6CF3/2020\|EPI_ISL_759914\|2020- | ···················· | ···················· | ····················· | ···················· | ···················· | ···················· |
| AB | 5904 | hCoV-19/Indonesia/JI-ITDua-998NTv/2020\|EPI_ISL_759 | ···················· | ···················· | ····················· | ···················· | ···················· | ···················· |
| AB | 5905 | hCoV-19/Indonesia/JI-ITDua-1006NTv/2020\|EPI_ISL_75 | ···················· | ···················· | ····················· | ···················· | ···················· | ···················· |
| AB | 5906 | hCoV-19/Indonesia/JI-ITDua-4134NTv/2020\|EPI_ISL_75 | ···················· | ···················· | ····················· | ···················· | ···················· | ···················· |
| AB | 5907 | hCoV-19/Indonesia/JI-ITDua-12323Nvv/2020\|EPI_ISL_7 | ···················· | ···················· | ····················· | ···················· | ···················· | ···················· |
| AB | 5908 | hCoV-19/Indonesia/JI-ITDua-6033NTv/2020\|EPI_ISL_75 | ···················· | ···················· | ····················· | ···················· | ···················· | ···················· |
| AB | 5909 | hCoV-19/Indonesia/JI-ITDua-2858NTv/2020\|EPI_ISL_75 | ···················· | ···················· | ····················· | ···················· | ···················· | ···················· |
| AB | 5910 | hCoV-19/Indonesia/JI-ITDua-1609Nv/2020\|EPI_ISL_759 | ···················· | ···················· | ····················· | ···················· | ···················· | ···················· |
| AB | 5911 | hCoV-19/Indonesia/JI-ITDua-28257NT/2020\|EPI_ISL_75 | ···················· | ···················· | ····················· | ···················· | ···················· | ···················· |
| AB | 5912 | hCoV-19/Norway/4367/2020\|EPI_ISL_759969\|2020-10-14 | ···················· | ···················· | ····················· | ···················· | ···················· | ···················· |
| AB | 5913 | hCoV-19/Norway/4368/2020\|EPI_ISL_759970\|2020-10-07 | ···················· | ···················· | ····················· | ···················· | ···················· | ···················· |
| AB | 5914 | hCoV-19/Norway/4371/2020\|EPI_ISL_759971\|2020-10-09 | ···················· | ···················· | ····················· | ···················· | ···················· | ···················· |
| AB | 5915 | hCoV-19/Norway/4372/2020\|EPI_ISL_759972\|2020-10-10 | ···················· | ···················· | ····················· | ···················· | ···················· | ···················· |
| AB | 5916 | hCoV-19/Norway/5824/2020\|EPI_ISL_759973\|2020-11-18 | ···················· | ···················· | ····················· | ···················· | ···················· | ···················· |
| AB | 5917 | hCoV-19/Norway/5845/2020\|EPI_ISL_759974\|2020-11-16 | ···················· | ···················· | ····················· | ···················· | ···················· | ···················· |
| AB | 5918 | hCoV-19/Norway/5850/2020\|EPI_ISL_759975\|2020-11-17 | ···················· | ···················· | ····················· | ···················· | ···················· | ···················· |
| AB | 5919 | hCoV-19/Norway/5896/2020\|EPI_ISL_759977\|2020-11-19 | ···················· | ···················· | ····················· | ···················· | ···················· | ···················· |
| AB | 5920 | hCoV-19/Norway/5897/2020\|EPI_ISL_759978\|2020-11-22 | ···················· | ···················· | ····················· | ···················· | ···················· | ···················· |
| AB | 5921 | hCoV-19/Norway/5898/2020\|EPI_ISL_759979\|2020-11-21 | ···················· | ·A·················· | ····················· | ···················· | ···················· | ··A················· |
| AB | 5922 | hCoV-19/Norway/5899/2020\|EPI_ISL_759980\|2020-11-22 | ···················· | ···················· | ····················· | ···················· | ···················· | ···················· |
| AB | 5923 | hCoV-19/Norway/5867/2020\|EPI_ISL_759981\|2020-11-23 | ···················· | ···················· | ····················· | ···················· | ···················· | ···················· |
| AB | 5924 | hCoV-19/Norway/6131/2020\|EPI_ISL_759982\|2020-11-28 | ···················· | ···················· | ····················· | ···················· | ···················· | ···················· |
| AB | 5925 | hCoV-19/Norway/6142/2020\|EPI_ISL_759983\|2020-11-29 | ···················· | ·A·················· | ····················· | ···················· | ···················· | ··A················· |
| AB | 5926 | hCoV-19/Norway/6388/2020\|EPI_ISL_759984\|2020-12-03 | ···················· | ·A·················· | ····················· | ···················· | ···················· | ··A················· |
| AB | 5927 | hCoV-19/Norway/6390/2020\|EPI_ISL_759985\|2020-12-02 | ···················· | ·A·················· | ····················· | ···················· | ···················· | ··A················· |
| AB | 5928 | hCoV-19/Norway/6421/2020\|EPI_ISL_759986\|2020-12-02 | ···················· | ·A·················· | ····················· | ···················· | ···················· | ··A················· |
| AB | 5929 | hCoV-19/Norway/6425/2020\|EPI_ISL_759988\|2020-12-01 | ···················· | ···················· | ····················· | ···················· | ······T············· | ···················· |
| AB | 5930 | hCoV-19/Norway/6435/2020\|EPI_ISL_759989\|2020-12-08 | ···················· | ···················· | ····················· | ···················· | ···················· | ···················· |
| AB | 5931 | hCoV-19/Norway/6458/2020\|EPI_ISL_759990\|2020-12-04 | ···················· | ···················· | ····················· | ···················· | ···················· | ···················· |
| AB | 5932 | hCoV-19/South Korea/KDCA0226/2020\|EPI_ISL_759991\|2 | ···················· | ···················· | ····················· | ···················· | ···················· | ···················· |
| AB | 5933 | hCoV-19/South Korea/KDCA0227/2020\|EPI_ISL_759992\|2 | ···················· | ···················· | ····················· | ···················· | ···················· | ···················· |
| AB | 5934 | hCoV-19/South Korea/KDCA0228/2020\|EPI_ISL_759993\|2 | ···················· | ···················· | ····················· | ···················· | ···················· | ···················· |
| AB | 5935 | hCoV-19/South Korea/KDCA0229/2020\|EPI_ISL_759994\|2 | ···················· | ···················· | ····················· | ···················· | ···················· | ···················· |
| AB | 5936 | hCoV-19/South Korea/KDCA0230/2020\|EPI_ISL_759995\|2 | ···················· | ···················· | ····················· | ···················· | ···················· | ···················· |
| AB | 5937 | hCoV-19/South Korea/KDCA0231/2020\|EPI_ISL_759996\|2 | ···················· | ···················· | ····················· | ···················· | ···················· | ··A················· |
| AB | 5938 | hCoV-19/South Korea/KDCA0232/2020\|EPI_ISL_759997\|2 | ···················· | ···················· | ····················· | ···················· | ···················· | ···················· |
| AB | 5939 | hCoV-19/South Korea/KDCA0233/2020\|EPI_ISL_759998\|2 | ···················· | ···················· | ····················· | ···················· | ···················· | ···················· |
| AB | 5940 | hCoV-19/South Korea/KDCA0234/2020\|EPI_ISL_759999\|2 | ···················· | ···················· | ····················· | ···················· | ···················· | ···················· |
| AB | 5941 | hCoV-19/England/ALDP-CB4DEA/2020\|EPI_ISL_760941\|20 | ···················· | ···················· | ····················· | ···················· | ···················· | ···················· |
| AB | 5942 | hCoV-19/England/MILK-CB2A8B/2020\|EPI_ISL_760943\|20 | ···················· | ···················· | ····················· | ···················· | ···················· | ···················· |
| AB | 5943 | hCoV-19/England/ALDP-CB47BF/2020\|EPI_ISL_760945\|20 | ···················· | ···················· | ····················· | ···················· | ···················· | ···················· |
| AB | 5944 | hCoV-19/England/ALDP-CB4DF9/2020\|EPI_ISL_760948\|20 | ···················· | ···················· | ····················· | ···················· | ···················· | ···················· |
| AB | 5945 | hCoV-19/England/ALDP-CB2687/2020\|EPI_ISL_760949\|20 | ···················· | ·A·················· | ····················· | ···················· | ···················· | ···················· |
| AB | 5946 | hCoV-19/England/ALDP-CB15D6/2020\|EPI_ISL_760952\|20 | ···················· | ···················· | ····················· | ···················· | ······T············· | ···················· |
| AB | 5947 | hCoV-19/England/MILK-CB1C68/2020\|EPI_ISL_760954\|20 | ···················· | ···················· | ····················· | ···················· | ···················· | ···················· |
| AB | 5948 | hCoV-19/England/MILK-C307AC/2020\|EPI_ISL_760959\|20 | ···················· | ···················· | ····················· | ···················· | ···················· | ···················· |
| AB | 5949 | hCoV-19/England/ALDP-CB3D90/2020\|EPI_ISL_760960\|20 | ···················· | ···················· | ····················· | ···················· | ···················· | ···················· |
| AB | 5950 | hCoV-19/England/MILK-CB0AD8/2020\|EPI_ISL_760961\|20 | ···················· | ·A·················· | ····················· | ···················· | ···················· | ··A················· |
| AB | 5951 | hCoV-19/England/MILK-C0BAB0/2020\|EPI_ISL_760963\|20 | ···················· | ···················· | ····················· | ···················· | ···················· | ···················· |
| AB | 5952 | hCoV-19/England/QEUH-CAD045/2020\|EPI_ISL_760964\|20 | ···················· | ···················· | ····················· | ···················· | ······T············· | ···················· |
| AB | 5953 | hCoV-19/England/ALDP-CB3D36/2020\|EPI_ISL_760967\|20 | ···················· | ···················· | ····················· | ···················· | ···················· | ···················· |
| AB | 5954 | hCoV-19/England/CAMC-C11A3F/2020\|EPI_ISL_760969\|20 | ···················· | ···················· | ····················· | ···················· | ·········T·········· | ···················· |
| AB | 5955 | hCoV-19/England/QEUH-CABDB9/2020\|EPI_ISL_760970\|20 | ···················· | ···················· | ····················· | ···················· | ···················· | ···················· |
| AB | 5956 | hCoV-19/England/ALDP-CB4E14/2020\|EPI_ISL_760971\|20 | ···················· | ···················· | ····················· | ···················· | ···················· | ···················· |
| AB | 5957 | hCoV-19/England/QEUH-CAD476/2020\|EPI_ISL_760974\|20 | ···················· | ···················· | ····················· | ···················· | ···················· | ···················· |
| AB | 5958 | hCoV-19/England/CAMC-CB3282/2020\|EPI_ISL_760976\|20 | ···················· | ·A·················· | ····················· | ···················· | ···················· | ··A················· |
| AB | 5959 | hCoV-19/England/ALDP-CB2757/2020\|EPI_ISL_760978\|20 | ···················· | ···················· | ····················· | ···················· | ···················· | ···················· |
| AB | 5960 | hCoV-19/England/ALDP-CB4719/2020\|EPI_ISL_760979\|20 | ···················· | ···················· | ····················· | ···················· | ···················· | ···················· |
| AB | 5961 | hCoV-19/England/ALDP-CB34B9/2020\|EPI_ISL_760982\|20 | ···················· | ···················· | ····················· | ···················· | ···················· | ···················· |
| AB | 5962 | hCoV-19/England/MILK-CB0953/2020\|EPI_ISL_760983\|20 | ···················· | ···················· | ····················· | ···················· | ···················· | ···················· |
| AB | 5963 | hCoV-19/England/QEUH-C5F464/2020\|EPI_ISL_760984\|20 | ···················· | ···················· | ····················· | ···················· | ···················· | ···················· |
| AB | 5964 | hCoV-19/England/QEUH-CB115A/2020\|EPI_ISL_760986\|20 | ···················· | ···················· | ····················· | ···················· | ···················· | ···················· |
| AB | 5965 | hCoV-19/England/MILK-CB14CA/2020\|EPI_ISL_760987\|20 | ···················· | ···················· | ····················· | ···················· | ···················· | ···················· |
| AB | 5966 | hCoV-19/England/CAMC-CB211D/2020\|EPI_ISL_760988\|20 | ···················· | ···················· | ····················· | ···················· | ···················· | ···················· |
| AB | 5967 | hCoV-19/England/ALDP-CAE6E8/2020\|EPI_ISL_760989\|20 | ···················· | ···················· | ····················· | ···················· | ···················· | ···················· |
| AB | 5968 | hCoV-19/England/QEUH-CAD090/2020\|EPI_ISL_760990\|20 | ···················· | ·A·················· | ····················· | ···················· | ···················· | ··A················· |
| AB | 5969 | hCoV-19/England/QEUH-CAD4EF/2020\|EPI_ISL_760992\|20 | ···················· | ···················· | ····················· | ···················· | ···················· | ···················· |
| AB | 5970 | hCoV-19/England/ALDP-CB4218/2020\|EPI_ISL_760994\|20 | ···················· | ···················· | ····················· | ···················· | ···················· | ···················· |
| AB | 5971 | hCoV-19/England/QEUH-CADD11/2020\|EPI_ISL_760995\|20 | ···················· | ···················· | ····················· | ···················· | ···················· | ···················· |
| AB | 5972 | hCoV-19/England/QEUH-CAA5EF/2020\|EPI_ISL_760996\|20 | ···················· | ···················· | ····················· | ···················· | ···················· | ···················· |
| AB | 5973 | hCoV-19/England/QEUH-CAD9C2/2020\|EPI_ISL_760999\|20 | ···················· | ···················· | ····················· | ···················· | ···················· | ···················· |
| AB | 5974 | hCoV-19/Netherlands/OV-EMC-121/2020\|EPI_ISL_763161 | ···················· | ···················· | ····················· | ···················· | ···················· | ···················· |
| AB | 5975 | hCoV-19/Netherlands/OV-EMC-123/2020\|EPI_ISL_763163 | ···················· | ···················· | ····················· | ···················· | ···················· | ···················· |
| AB | 5976 | hCoV-19/Netherlands/OV-EMC-124/2020\|EPI_ISL_763164 | ···················· | ···················· | ····················· | ···················· | ···················· | ···················· |
| AB | 5977 | hCoV-19/Netherlands/OV-EMC-125/2020\|EPI_ISL_763165 | ···················· | ······A············· | ····················· | ···················· | ···················· | ···················· |

| | | | | | | | |
|---|---|---|---|---|---|---|---|
| AB | 5978 hCoV-19/Netherlands/OV-EMC-126/2020\|EPI_ISL_763166 | ···················· | ···················· | ····················· | ···················· | ···················· | ···················· |
| AB | 5979 hCoV-19/Netherlands/OV-EMC-128/2020\|EPI_ISL_763167 | ···················· | ···················· | ····················· | ···················· | ···················· | ···················· |
| AB | 5980 hCoV-19/Netherlands/OV-EMC-129/2020\|EPI_ISL_763168 | ···················· | ···················· | ····················· | ···················· | ···················· | ···················· |
| AB | 5981 hCoV-19/Netherlands/OV-EMC-130/2020\|EPI_ISL_763169 | ···················· | ···················· | ····················· | ···················· | ···················· | ···················· |
| AB | 5982 hCoV-19/Netherlands/OV-EMC-131/2020\|EPI_ISL_763170 | ···················· | ···················· | ····················· | ···················· | ···················· | ···················· |
| AB | 5983 hCoV-19/Netherlands/OV-EMC-132/2020\|EPI_ISL_763171 | ···················· | ···················· | ····················· | ···················· | ···················· | ···················· |
| AB | 5984 hCoV-19/Netherlands/un-EMC-819/2020\|EPI_ISL_763172 | ···················· | ···················· | ····················· | ···················· | ···················· | ···················· |
| AB | 5985 hCoV-19/Netherlands/un-EMC-821/2020\|EPI_ISL_763174 | ···················· | ···················· | ····················· | ···················· | ···················· | ···················· |
| AB | 5986 hCoV-19/Netherlands/un-EMC-822/2020\|EPI_ISL_763175 | ···················· | ···················· | ····················· | ···················· | ···················· | ···················· |
| AB | 5987 hCoV-19/Netherlands/un-EMC-823/2020\|EPI_ISL_763176 | ···················· | ···················· | ····················· | ···················· | ···················· | ···················· |
| AB | 5988 hCoV-19/Netherlands/un-EMC-824/2020\|EPI_ISL_763177 | ···················· | ···················· | ····················· | ···················· | ···················· | ···················· |
| AB | 5989 hCoV-19/Netherlands/un-EMC-825/2020\|EPI_ISL_763178 | ···················· | ···················· | ····················· | ···················· | ···················· | ···················· |
| AB | 5990 hCoV-19/Netherlands/un-EMC-826/2020\|EPI_ISL_763179 | ···················· | ···················· | ····················· | ···················· | ···················· | ···················· |
| AB | 5991 hCoV-19/Netherlands/ZH-EMC-1146/2020\|EPI_ISL_76322 | ···················· | ···················· | ····················· | ···················· | ···················· | ···················· |
| AB | 5992 hCoV-19/England/NOTT-121787/2020\|EPI_ISL_764494\|20 | ···················· | ···················· | ····················· | ···················· | ···················· | ···················· |
| AB | 5993 hCoV-19/England/NOTT-121801/2020\|EPI_ISL_764495\|20 | ···················· | ···················· | ····················· | ···················· | ···················· | ···················· |
| AB | 5994 hCoV-19/England/NOTT-121803/2020\|EPI_ISL_764496\|20 | ···················· | ···················· | ····················· | ···················· | ···················· | ···················· |
| AB | 5995 hCoV-19/England/NOTT-121805/2020\|EPI_ISL_764497\|20 | ···················· | ···················· | ····················· | ···················· | ···················· | ···················· |
| AB | 5996 hCoV-19/England/NOTT-121813/2020\|EPI_ISL_764498\|20 | ···················· | ···················· | ····················· | ···················· | ···················· | ···················· |
| AB | 5997 hCoV-19/England/NOTT-121817/2020\|EPI_ISL_764499\|20 | ···················· | ···················· | ····················· | ···················· | ···················· | ···················· |
| AB | 5998 hCoV-19/England/NOTT-121822/2020\|EPI_ISL_764500\|20 | ···················· | ···················· | ····················· | ···················· | ···················· | ···················· |
| AB | 5999 hCoV-19/England/NOTT-121826/2020\|EPI_ISL_764501\|20 | ···················· | ·A·······A·········· | ····················· | ···················· | ···················· | ···················· |
| AB | 6000 hCoV-19/Wales/OXON-F40F26/2020\|EPI_ISL_764555\|2020 | ···················· | ···················· | ····················· | ···················· | ···················· | ···················· |
| AB | 6001 hCoV-19/USA/MA-MGH-03353/2020\|EPI_ISL_765800\|2020- | ···················· | ···················· | ····················· | ···················· | ···················· | ···················· |
| AB | 6002 hCoV-19/USA/MA-MGH-03357/2020\|EPI_ISL_765801\|2020- | ···················· | ···················· | ····················· | ···················· | ···················· | ···················· |
| AB | 6003 hCoV-19/USA/MA-MGH-03360/2020\|EPI_ISL_765802\|2020- | ···················· | ···················· | ····················· | ···················· | ···················· | ···················· |
| AB | 6004 hCoV-19/USA/MA-MGH-03361/2020\|EPI_ISL_765803\|2020- | ···················· | ···················· | ····················· | ···················· | ···················· | ···················· |
| AB | 6005 hCoV-19/USA/MA-MGH-03365/2020\|EPI_ISL_765805\|2020- | ···················· | ···················· | ····················· | ···················· | ···················· | ···················· |
| AB | 6006 hCoV-19/USA/MA-MGH-03368/2020\|EPI_ISL_765806\|2020- | ···················· | ···················· | ····················· | ···················· | ···················· | ···················· |
| AB | 6007 hCoV-19/USA/MA-MGH-03370/2020\|EPI_ISL_765807\|2020- | ···················· | ·A·················· | ····················· | ···················· | ···················· | ···················· |
| AB | 6008 hCoV-19/USA/MA-MGH-03372/2020\|EPI_ISL_765808\|2020- | ···················· | ···················· | ····················· | ···················· | ···················· | ···················· |
| AB | 6009 hCoV-19/USA/MA-MGH-03374/2020\|EPI_ISL_765809\|2020- | ···················· | ···················· | ····················· | ···················· | ···················· | ···················· |
| AB | 6010 hCoV-19/USA/MA-MGH-03375/2020\|EPI_ISL_765810\|2020- | ···················· | ···················· | ····················· | ···················· | ···················· | ···················· |
| AB | 6011 hCoV-19/USA/MA-MGH-03384/2020\|EPI_ISL_765811\|2020- | ···················· | ·A·················· | ····················· | ···················· | ···················· | ···················· |
| AB | 6012 hCoV-19/USA/MA-MGH-03386/2020\|EPI_ISL_765812\|2020- | ···················· | ···················· | ····················· | ···················· | ···················· | ···················· |
| AB | 6013 hCoV-19/USA/MA-MGH-03389/2020\|EPI_ISL_765813\|2020- | ···················· | ···················· | ····················· | ···················· | ···················· | ···················· |
| AB | 6014 hCoV-19/USA/MA-MGH-03393/2020\|EPI_ISL_765814\|2020- | ···················· | ···················· | ····················· | ···················· | ···················· | ···················· |
| AB | 6015 hCoV-19/USA/MA-MGH-03396/2020\|EPI_ISL_765815\|2020- | ···················· | ···················· | ····················· | ···················· | ···················· | ···················· |
| AB | 6016 hCoV-19/USA/MA-MGH-03398/2020\|EPI_ISL_765816\|2020- | ···················· | ···················· | ····················· | ···················· | ···················· | ···················· |
| AB | 6017 hCoV-19/USA/MA-MGH-03399/2020\|EPI_ISL_765817\|2020- | ···················· | ···················· | ····················· | ···················· | ···················· | ···················· |
| AB | 6018 hCoV-19/USA/MA-MGH-03401/2020\|EPI_ISL_765819\|2020- | ···················· | ···················· | ····················· | ···················· | ···················· | ···················· |
| AB | 6019 hCoV-19/USA/MA-MGH-03402/2020\|EPI_ISL_765820\|2020- | ···················· | ···················· | ····················· | ···················· | ···················· | ···················· |
| AB | 6020 hCoV-19/USA/MA-MGH-03403/2020\|EPI_ISL_765821\|2020- | ···················· | ···················· | ····················· | ···················· | ···················· | ···················· |
| AB | 6021 hCoV-19/USA/MA-MGH-03405/2020\|EPI_ISL_765822\|2020- | ···················· | ···················· | ····················· | ···················· | ···················· | ···················· |
| AB | 6022 hCoV-19/USA/MA-MGH-03407/2020\|EPI_ISL_765823\|2020- | ···················· | ···················· | ····················· | ···················· | ···················· | ···················· |
| AB | 6023 hCoV-19/USA/MA-MGH-03414/2020\|EPI_ISL_765824\|2020- | ···················· | ···················· | ····················· | ···················· | ···················· | ···················· |
| AB | 6024 hCoV-19/USA/MA-MGH-03415/2020\|EPI_ISL_765825\|2020- | ···················· | ···················· | ····················· | ···················· | ···················· | ···················· |
| AB | 6025 hCoV-19/USA/MA-MGH-03416/2020\|EPI_ISL_765826\|2020- | ···················· | ···················· | ····················· | ···················· | ···················· | ···················· |
| AB | 6026 hCoV-19/USA/MA-MGH-03417/2020\|EPI_ISL_765827\|2020- | ···················· | ···················· | ····················· | ···················· | ···················· | ···················· |
| AB | 6027 hCoV-19/USA/MA-MGH-03418/2020\|EPI_ISL_765828\|2020- | ···················· | ···················· | ····················· | ···················· | ···················· | ···················· |
| AB | 6028 hCoV-19/USA/MA-MGH-03419/2020\|EPI_ISL_765829\|2020- | ···················· | ···················· | ····················· | ···················· | ···················· | ···················· |
| AB | 6029 hCoV-19/USA/MA-MGH-03421/2020\|EPI_ISL_765830\|2020- | ···················· | ···················· | ····················· | ···················· | ···················· | ···················· |
| AB | 6030 hCoV-19/USA/MA-MGH-03424/2020\|EPI_ISL_765831\|2020- | ···················· | ···················· | ····················· | ···················· | ···················· | ···················· |
| AB | 6031 hCoV-19/USA/MA-MGH-03426/2020\|EPI_ISL_765832\|2020- | ···················· | ···················· | ····················· | ···················· | ···················· | ···················· |
| AB | 6032 hCoV-19/USA/MA-MGH-03564/2020\|EPI_ISL_765886\|2020- | ···················· | ·A·················· | ····················· | ···················· | ···················· | ···················· |
| AB | 6033 hCoV-19/Switzerland/BE-ETHZ-420291/2020\|EPI_ISL_76 | ···················· | ···················· | ····················· | ···················· | ···················· | ···················· |
| AB | 6034 hCoV-19/Switzerland/BE-ETHZ-420312/2020\|EPI_ISL_76 | ···················· | ···················· | ····················· | ···················· | ··············K····· | ···················· |
| AB | 6035 hCoV-19/Switzerland/BE-ETHZ-420040/2020\|EPI_ISL_76 | ···················· | ···················· | ····················· | ···················· | ···················· | ···················· |
| AB | 6036 hCoV-19/Switzerland/BE-ETHZ-420681/2020\|EPI_ISL_76 | ···················· | ···················· | ····················· | ···················· | ···················· | ···················· |
| AB | 6037 hCoV-19/Switzerland/BE-ETHZ-420683/2020\|EPI_ISL_76 | ···················· | ···················· | ····················· | ···················· | ···················· | ···················· |
| AB | 6038 hCoV-19/Switzerland/BE-ETHZ-420625/2020\|EPI_ISL_76 | ···················· | ···················· | ····················· | ···················· | ···················· | ···················· |
| AB | 6041 hCoV-19/Switzerland/BE-ETHZ-420572/2020\|EPI_ISL_76 | ···················· | ···················· | ····················· | ···················· | ···················· | ···················· |
| AB | 6044 hCoV-19/Switzerland/BL-ETHZ-420702/2020\|EPI_ISL_76 | ···················· | ·········R·········· | ····················· | ···················· | ···················· | ···················· |
| AB | 6045 hCoV-19/Switzerland/BL-ETHZ-420204/2020\|EPI_ISL_76 | ···················· | ···················· | ····················· | ···················· | ···················· | ···················· |
| AB | 6048 hCoV-19/England/MILK-C60BC5/2020\|EPI_ISL_769301\|20 | ···················· | ···················· | ····················· | ···················· | ···················· | ···················· |
| AB | 6049 hCoV-19/England/MILK-C63290/2020\|EPI_ISL_769305\|20 | ···················· | ···················· | ····················· | ···················· | ···················· | ···················· |
| AB | 6050 hCoV-19/England/QEUH-C5BFFE/2020\|EPI_ISL_769310\|20 | ···················· | ···················· | ····················· | ···················· | ···················· | ···················· |
| AB | 6051 hCoV-19/England/QEUH-C5BAFD/2020\|EPI_ISL_769314\|20 | ···················· | ···················· | ····················· | ···················· | ···················· | ···················· |
| AB | 6052 hCoV-19/England/MILK-C618DC/2020\|EPI_ISL_769315\|20 | ···················· | ···················· | ····················· | ···················· | ···················· | ···················· |
| AB | 6053 hCoV-19/England/QEUH-C5BC06/2020\|EPI_ISL_769316\|20 | ···················· | ···················· | ····················· | ···················· | ···················· | ···················· |
| AB | 6054 hCoV-19/England/MILK-C5FF45/2020\|EPI_ISL_769318\|20 | ···················· | ···················· | ····················· | ···················· | ···················· | ···················· |
| AB | 6055 hCoV-19/England/QEUH-C5B978/2020\|EPI_ISL_769319\|20 | ···················· | ···················· | ····················· | ···················· | ···················· | ···················· |
| AB | 6056 hCoV-19/England/QEUH-C5CA83/2020\|EPI_ISL_769321\|20 | ···················· | ···················· | ····················· | ···················· | ···················· | ···················· |
| AB | 6057 hCoV-19/England/QEUH-C5D5BE/2020\|EPI_ISL_769322\|20 | ···················· | ···················· | ····················· | ···················· | ···················· | ···················· |
| AB | 6058 hCoV-19/England/QEUH-C5C43A/2020\|EPI_ISL_769323\|20 | ···················· | ···················· | ····················· | ···················· | ···················· | ···················· |
| AB | 6059 hCoV-19/England/MILK-C60F6F/2020\|EPI_ISL_769324\|20 | ···················· | ···················· | ····················· | ···················· | ···················· | ···················· |
| AB | 6060 hCoV-19/England/MILK-C61405/2020\|EPI_ISL_769326\|20 | ···················· | ···················· | ····················· | ···················· | ···················· | ···················· |
| AB | 6061 hCoV-19/England/QEUH-C5BF58/2020\|EPI_ISL_769330\|20 | ···················· | ···················· | ····················· | ···················· | ···················· | ···················· |
| AB | 6062 hCoV-19/England/MILK-C60F50/2020\|EPI_ISL_769332\|20 | ···················· | ···················· | ····················· | ···················· | ···················· | ···················· |
| AB | 6063 hCoV-19/England/QEUH-C5B9E1/2020\|EPI_ISL_769334\|20 | ···················· | ···················· | ····················· | ···················· | ···················· | ···················· |

| | | | | | | | | |
|---|---|---|---|---|---|---|---|---|
| AB | 6064 | hCoV-19/England/MILK-C60C86/2020\|EPI_ISL_769337\|20 | ···················· | ···················· | ····················· | ···················· | ···················· | ···A················ |
| AB | 6065 | hCoV-19/England/MILK-C61450/2020\|EPI_ISL_769340\|20 | ···················· | ···················· | ····················· | ···················· | ···················· | ···A················ |
| AB | 6066 | hCoV-19/England/MILK-C60CB3/2020\|EPI_ISL_769342\|20 | ···················· | ···················· | ····················· | ···················· | ···················· | ···A················ |
| AB | 6067 | hCoV-19/England/MILK-C618FA/2020\|EPI_ISL_769347\|20 | ···················· | ···················· | ····················· | ···················· | ······T············· | ···················· |
| AB | 6068 | hCoV-19/England/QEUH-C5B9A5/2020\|EPI_ISL_769348\|20 | ···················· | ···················· | ····················· | ···················· | ···················· | ···················· |
| AB | 6069 | hCoV-19/England/MILK-C609CB/2020\|EPI_ISL_769350\|20 | ···················· | ···················· | ····················· | ···················· | ···················· | ···················· |
| AB | 6070 | hCoV-19/England/QEUH-C5BD9A/2020\|EPI_ISL_769352\|20 | ···················· | ···················· | ····················· | ···················· | ···················· | ···················· |
| AB | 6071 | hCoV-19/England/MILK-C61441/2020\|EPI_ISL_769354\|20 | ···················· | ···················· | ····················· | ···················· | ···················· | ···A················ |
| AB | 6072 | hCoV-19/England/QEUH-C5B9D2/2020\|EPI_ISL_769355\|20 | ···················· | ···················· | ····················· | ···················· | ···················· | ···················· |
| AB | 6073 | hCoV-19/England/MILK-C60C95/2020\|EPI_ISL_769357\|20 | ···················· | ···················· | ····················· | ···················· | ···················· | ···················· |
| AB | 6074 | hCoV-19/England/QEUH-C5D8F1/2020\|EPI_ISL_769360\|20 | ···················· | ···················· | ····················· | ···················· | ···················· | ···················· |
| AB | 6075 | hCoV-19/England/MILK-C5FFBE/2020\|EPI_ISL_769364\|20 | ···················· | ···················· | ····················· | ···················· | ···················· | ···················· |
| AB | 6076 | hCoV-19/England/QEUH-C5BFEF/2020\|EPI_ISL_769365\|20 | ···················· | ···················· | ····················· | ···················· | ···················· | ···················· |
| AB | 6077 | hCoV-19/England/QEUH-C5BB45/2020\|EPI_ISL_769367\|20 | ···················· | ···················· | ····················· | ···················· | ···················· | ···················· |
| AB | 6078 | hCoV-19/England/MILK-C63324/2020\|EPI_ISL_769368\|20 | ···················· | ···················· | ····················· | ···················· | ···················· | ···················· |
| AB | 6079 | hCoV-19/England/MILK-C6199D/2020\|EPI_ISL_769373\|20 | ···················· | ···················· | ····················· | ···················· | ···················· | ···················· |
| AB | 6080 | hCoV-19/England/QEUH-C5C036/2020\|EPI_ISL_769374\|20 | ···················· | ······A············· | ····················· | ···················· | ···················· | ···················· |
| AB | 6081 | hCoV-19/England/MILK-C60E35/2020\|EPI_ISL_769375\|20 | ···················· | ···················· | ····················· | ···················· | ···················· | ···················· |
| AB | 6082 | hCoV-19/England/MILK-C60DFC/2020\|EPI_ISL_769376\|20 | ···················· | ···················· | ····················· | ···················· | ···················· | ···················· |
| AB | 6083 | hCoV-19/England/MILK-C60CFF/2020\|EPI_ISL_769378\|20 | ···················· | ···················· | ····················· | ···················· | ···················· | ···················· |
| AB | 6084 | hCoV-19/England/QEUH-C5C018/2020\|EPI_ISL_769385\|20 | ···················· | ···················· | ····················· | ···················· | ···················· | ···················· |
| AB | 6085 | hCoV-19/England/QEUH-C5BE79/2020\|EPI_ISL_769388\|20 | ···················· | ···················· | ····················· | ···················· | ···················· | ···················· |
| AB | 6086 | hCoV-19/England/QEUH-C5BF76/2020\|EPI_ISL_769389\|20 | ···················· | ···················· | ····················· | ···················· | ···················· | ···················· |
| AB | 6087 | hCoV-19/England/QEUH-C5BF85/2020\|EPI_ISL_769391\|20 | ···················· | ···················· | ····················· | ···················· | ···················· | ···················· |
| AB | 6088 | hCoV-19/England/MILK-C6149C/2020\|EPI_ISL_769393\|20 | ···················· | ···················· | ····················· | ···················· | ···················· | ···················· |
| AB | 6089 | hCoV-19/England/QEUH-C5CAA1/2020\|EPI_ISL_769394\|20 | ···················· | ···················· | ····················· | ···················· | ···················· | ···················· |
| AB | 6090 | hCoV-19/England/QEUH-C5C379/2020\|EPI_ISL_769399\|20 | ···················· | ···················· | ····················· | ···················· | ···················· | ···················· |
| AB | 6091 | hCoV-19/Canada/ON-PHL-20-03635/2020\|EPI_ISL_775139 | ···················· | ···················· | ····················· | ···················· | ···················· | ···················· |
| AB | 6092 | hCoV-19/Canada/ON-PHL-20-03636/2020\|EPI_ISL_775140 | ···················· | ···················· | ····················· | ···················· | ···················· | ···················· |
| AB | 6093 | hCoV-19/Canada/ON-PHL-20-03637/2020\|EPI_ISL_775141 | ···················· | ···················· | ····················· | ···················· | ···················· | ···················· |
| AB | 6094 | hCoV-19/Canada/ON-PHL-20-03638/2020\|EPI_ISL_775142 | ···················· | ···················· | ····················· | ···················· | ···················· | ···················· |
| AB | 6095 | hCoV-19/Canada/ON-PHL-20-03639/2020\|EPI_ISL_775143 | ···················· | ·A·················· | ····················· | ···················· | ···················· | ···A················ |
| AB | 6096 | hCoV-19/Canada/ON-PHL-20-03640/2020\|EPI_ISL_775144 | ···················· | ·A·················· | ····················· | ···················· | ···················· | ···A················ |
| AB | 6097 | hCoV-19/Canada/ON-PHL-20-03641/2020\|EPI_ISL_775145 | ···················· | ·A·················· | ····················· | ···················· | ···················· | ···A················ |
| AB | 6098 | hCoV-19/Canada/ON-PHL-20-03642/2020\|EPI_ISL_775146 | ···················· | ·A·················· | ····················· | ···················· | ···················· | ···A················ |
| AB | 6099 | hCoV-19/Canada/ON-PHL-20-03643/2020\|EPI_ISL_775147 | ···················· | ···················· | ····················· | ···················· | ···················· | ···················· |
| AB | 6100 | hCoV-19/Canada/ON-PHL-20-03644/2020\|EPI_ISL_775148 | ···················· | ·A·················· | ····················· | ···················· | ···················· | ···A················ |
| AB | 6101 | hCoV-19/Canada/ON-PHL-20-03645/2020\|EPI_ISL_775149 | ···················· | ·A·················· | ····················· | ···················· | ···················· | ···A················ |
| AB | 6102 | hCoV-19/Canada/ON-PHL-20-03646/2020\|EPI_ISL_775150 | ···················· | ·A·················· | ····················· | ···················· | ···················· | ···A················ |
| AB | 6103 | hCoV-19/Canada/ON-PHL-20-03647/2020\|EPI_ISL_775151 | ···················· | ······A············· | ····················· | ···················· | ···················· | ···················· |
| AB | 6104 | hCoV-19/Canada/ON-PHL-20-03649/2020\|EPI_ISL_775152 | ···················· | ·A·················· | ····················· | ···················· | ···················· | ···A················ |
| AB | 6105 | hCoV-19/Canada/ON-PHL-20-03667/2020\|EPI_ISL_775153 | ···················· | ·A·················· | ····················· | ···················· | ···················· | ···A················ |
| AB | 6106 | hCoV-19/Canada/ON-PHL-20-03668/2020\|EPI_ISL_775154 | ···················· | ·A·················· | ····················· | ···················· | ···················· | ···A················ |
| AB | 6107 | hCoV-19/Canada/ON-PHL-20-03669/2020\|EPI_ISL_775155 | ···················· | ·A·················· | ····················· | ···················· | ···················· | ···A················ |
| AB | 6108 | hCoV-19/Canada/ON-PHL-20-03670/2020\|EPI_ISL_775156 | ···················· | ·A·················· | ····················· | ···················· | ···················· | ···A················ |
| AB | 6109 | hCoV-19/Canada/ON-PHL-20-03671/2020\|EPI_ISL_775157 | ···················· | ·········A·········· | ····················· | ···················· | ···················· | ···················· |
| AB | 6110 | hCoV-19/Canada/ON-PHL-20-03672/2020\|EPI_ISL_775158 | ···················· | ···················· | ····················· | ···················· | ···················· | ···················· |
| AB | 6111 | hCoV-19/Canada/ON-PHL-20-03675/2020\|EPI_ISL_775159 | ···················· | ···················· | ····················· | ···················· | ···················· | ···················· |
| AB | 6112 | hCoV-19/Canada/ON-PHL-20-03733/2020\|EPI_ISL_775206 | ···················· | ···················· | ····················· | ···················· | ···················· | ···················· |
| AB | 6113 | hCoV-19/Canada/ON-PHL-20-03737/2020\|EPI_ISL_775207 | ···················· | ···················· | ····················· | ···················· | ···················· | ···················· |
| AB | 6114 | hCoV-19/Canada/ON-PHL-20-03740/2020\|EPI_ISL_775208 | ···················· | ···················· | ····················· | ···················· | NN·················· | ···················· |
| AB | 6115 | hCoV-19/Canada/ON-PHL-20-03741/2020\|EPI_ISL_775209 | ···················· | ···················· | ····················· | ···················· | ···················· | ···················· |
| AB | 6116 | hCoV-19/Germany/HH-hpi-p2395/2020\|EPI_ISL_776470\|2 | ···················· | ···················· | ····················· | ···················· | ···················· | ···················· |
| AB | 6117 | hCoV-19/Germany/HH-hpi-p2396/2020\|EPI_ISL_776471\|2 | ···················· | ···················· | ····················· | ···················· | ···················· | ···················· |
| AB | 6118 | hCoV-19/Germany/HH-hpi-p2397/2020\|EPI_ISL_776472\|2 | ···················· | ···················· | ····················· | ···················· | ···················· | ···················· |
| AB | 6119 | hCoV-19/Germany/HH-hpi-p2398/2020\|EPI_ISL_776473\|2 | ···················· | ···················· | ····················· | ···················· | ···················· | ···················· |
| AB | 6120 | hCoV-19/Germany/HH-hpi-p2401/2020\|EPI_ISL_776474\|2 | ···················· | ···················· | ····················· | ···················· | ···················· | ···················· |
| AB | 6121 | hCoV-19/Germany/HH-hpi-p2402/2020\|EPI_ISL_776475\|2 | ···················· | ···················· | ····················· | ···················· | ···················· | ···················· |
| AB | 6122 | hCoV-19/Germany/HH-hpi-p2403/2020\|EPI_ISL_776476\|2 | ···················· | ···················· | ····················· | ···················· | ···················· | ···················· |
| AB | 6123 | hCoV-19/Germany/HH-hpi-p2404/2020\|EPI_ISL_776477\|2 | ···················· | ···················· | ····················· | ···················· | ···················· | ···················· |
| AB | 6124 | hCoV-19/Germany/HH-hpi-p2405/2020\|EPI_ISL_776478\|2 | ···················· | ···················· | ····················· | ···················· | ···················· | ···················· |
| AB | 6125 | hCoV-19/Germany/HH-hpi-p2406/2020\|EPI_ISL_776479\|2 | ···················· | ···················· | ····················· | ···················· | ···················· | ···················· |
| AB | 6126 | hCoV-19/Germany/HH-hpi-p2407/2020\|EPI_ISL_776480\|2 | ···················· | ···················· | ····················· | ···················· | ···················· | ···················· |
| AB | 6127 | hCoV-19/Germany/HH-hpi-p2408/2020\|EPI_ISL_776481\|2 | ···················· | ···················· | ····················· | ···················· | ···················· | ···················· |
| AB | 6128 | hCoV-19/Germany/HH-hpi-p2410/2020\|EPI_ISL_776482\|2 | ···················· | ···················· | ····················· | ···················· | ···················· | ···················· |
| AB | 6129 | hCoV-19/Germany/HH-hpi-p2412/2020\|EPI_ISL_776483\|2 | ···················· | ···················· | ····················· | ···················· | ···················· | ···················· |
| AB | 6130 | hCoV-19/Germany/HH-hpi-p2413/2020\|EPI_ISL_776484\|2 | ···················· | ···················· | ····················· | ···················· | ···················· | ···················· |
| AB | 6132 | hCoV-19/Germany/HH-hpi-p2416/2020\|EPI_ISL_776486\|2 | ···················· | ···················· | ····················· | ···················· | ···················· | ···················· |
| AB | 6133 | hCoV-19/Germany/HH-hpi-p2417/2020\|EPI_ISL_776487\|2 | ···················· | ···················· | ····················· | ···················· | ···················· | ···················· |
| AB | 6134 | hCoV-19/Germany/HH-hpi-p2419/2020\|EPI_ISL_776488\|2 | ···················· | ···················· | ····················· | ···················· | ···················· | ···················· |
| AB | 6135 | hCoV-19/Germany/HH-hpi-p2420/2020\|EPI_ISL_776489\|2 | ···················· | ···················· | ····················· | ···················· | ···················· | ···················· |
| AB | 6136 | hCoV-19/Germany/HH-hpi-p2478/2020\|EPI_ISL_776536\|2 | ···················· | ···················· | ····················· | ···················· | ···················· | ···················· |
| AB | 6138 | hCoV-19/Germany/HH-hpi-p2480/2020\|EPI_ISL_776538\|2 | ···················· | ···················· | ····················· | ···················· | ···················· | ···················· |
| AB | 6139 | hCoV-19/Germany/HH-hpi-p2482/2020\|EPI_ISL_776539\|2 | ···················· | ···················· | ····················· | ···················· | ···················· | ···················· |
| AB | 6140 | hCoV-19/England/CAMC-CB7910/2020\|EPI_ISL_777800\|20 | ···················· | ···················· | ····················· | ···················· | ···················· | ···················· |
| AB | 6141 | hCoV-19/England/CAMC-CB894C/2020\|EPI_ISL_777801\|20 | ···················· | ···················· | ····················· | ···················· | ······T············· | ···················· |
| AB | 6142 | hCoV-19/England/CAMC-CB75A3/2020\|EPI_ISL_777803\|20 | ···················· | ···················· | ····················· | ···················· | ···················· | ···················· |
| AB | 6143 | hCoV-19/England/CAMC-CB7D23/2020\|EPI_ISL_777811\|20 | ···················· | ···················· | ····················· | ···················· | ···················· | ···················· |
| AB | 6144 | hCoV-19/England/CAMC-CB7F5A/2020\|EPI_ISL_777869\|20 | ···················· | ···················· | ····················· | ···················· | ···················· | ···················· |
| AB | 6145 | hCoV-19/Netherlands/ZH-EMC-1121/2020\|EPI_ISL_76320 | ···················· | ···················· | ····················· | ···················· | ···················· | ···················· |

| | | | | | | | | |
|---|---|---|---|---|---|---|---|---|
| AB | 6146 | hCoV-19/Netherlands/ZH-EMC-1123/2020\|EPI_ISL_76320 | ···················· | ···················· | ···················· | ···················· | ······T············· | ···················· |
| AB | 6147 | hCoV-19/Netherlands/ZH-EMC-1124/2020\|EPI_ISL_76320 | ···················· | ···················· | ···················· | ···················· | ···················· | ···················· |
| AB | 6148 | hCoV-19/Netherlands/ZH-EMC-1138/2020\|EPI_ISL_76321 | ···················· | ···················· | ···················· | ···················· | ···················· | ···················· |
| AB | 6149 | hCoV-19/Netherlands/ZH-EMC-1139/2020\|EPI_ISL_76321 | ···················· | ···················· | ···················· | ···················· | ···················· | ···················· |
| AB | 6150 | hCoV-19/Netherlands/ZH-EMC-1140/2020\|EPI_ISL_76321 | ···················· | ····A··············· | ···················· | ···················· | ···················· | ···················· |
| AB | 6151 | hCoV-19/Netherlands/ZH-EMC-1141/2020\|EPI_ISL_76321 | ···················· | ···················· | ···················· | ···················· | ···················· | ···················· |
| AB | 6152 | hCoV-19/Netherlands/ZH-EMC-1142/2020\|EPI_ISL_76321 | ···················· | ···················· | ···················· | ···················· | ···················· | ···················· |
| AB | 6153 | hCoV-19/Netherlands/ZH-EMC-1143/2020\|EPI_ISL_76322 | ···················· | ···················· | ···················· | ···················· | ···················· | ···················· |
| AB | 6154 | hCoV-19/Netherlands/ZH-EMC-1144/2020\|EPI_ISL_76322 | ···················· | ···················· | ···················· | ···················· | ···················· | ···················· |
| AB | 6155 | hCoV-19/Netherlands/ZH-EMC-1147/2020\|EPI_ISL_76322 | ···················· | ···················· | ···················· | ···················· | ···················· | ···················· |
| AB | 6156 | hCoV-19/Netherlands/ZH-EMC-1149/2020\|EPI_ISL_76322 | ···················· | ···················· | ···················· | ···················· | ···················· | ···················· |
| AB | 6157 | hCoV-19/Netherlands/ZH-EMC-1150/2020\|EPI_ISL_76322 | ···················· | ···················· | ···················· | ···················· | ···················· | ···················· |
| AB | 6158 | hCoV-19/Netherlands/ZH-EMC-1151/2020\|EPI_ISL_76322 | ···················· | ···················· | ···················· | ···················· | ···················· | ···················· |
| AB | 6159 | hCoV-19/Netherlands/ZH-EMC-1154/2020\|EPI_ISL_76322 | ···················· | ···················· | ···················· | ···················· | ···················· | ···················· |
| AB | 6160 | hCoV-19/Netherlands/ZH-EMC-1155/2020\|EPI_ISL_76323 | ···················· | ···················· | ···················· | ···················· | ···················· | ···················· |
| AB | 6161 | hCoV-19/Netherlands/ZH-EMC-1156/2020\|EPI_ISL_76323 | ···················· | ···················· | ···················· | ···················· | ···················· | ···················· |
| AB | 6162 | hCoV-19/Netherlands/ZH-EMC-1158/2020\|EPI_ISL_76323 | ···················· | ···················· | ···················· | ···················· | ···················· | ···················· |
| AB | 6163 | hCoV-19/Netherlands/ZH-EMC-1160/2020\|EPI_ISL_76323 | ···················· | ···················· | ···················· | ···················· | ···················· | ···················· |
| AB | 6164 | hCoV-19/Netherlands/ZH-EMC-1161/2020\|EPI_ISL_76323 | ···················· | ···················· | ···················· | ···················· | ···················· | ···················· |
| AB | 6165 | hCoV-19/Netherlands/ZH-EMC-1162/2020\|EPI_ISL_76323 | ···················· | ···················· | ···················· | ···················· | ···················· | ···················· |
| AB | 6166 | hCoV-19/Netherlands/ZH-EMC-1164/2020\|EPI_ISL_76323 | ···················· | ···················· | ···················· | ···················· | ···················· | ···················· |
| AB | 6167 | hCoV-19/Netherlands/ZH-EMC-1166/2020\|EPI_ISL_76324 | ···················· | ···················· | ···················· | ···················· | ······T············· | ···················· |
| AB | 6168 | hCoV-19/Netherlands/ZH-EMC-1168/2020\|EPI_ISL_76324 | ···················· | ···················· | ···················· | ···················· | ···················· | ···················· |
| AB | 6169 | hCoV-19/Netherlands/ZH-EMC-1169/2020\|EPI_ISL_76324 | ···················· | ···················· | ···················· | ···················· | ···················· | ···················· |
| AB | 6170 | hCoV-19/Netherlands/ZH-EMC-1170/2020\|EPI_ISL_76324 | ···················· | ···················· | ···················· | ···················· | ···················· | ···················· |
| AB | 6171 | hCoV-19/Netherlands/ZH-EMC-1171/2020\|EPI_ISL_76324 | ···················· | ···················· | ···················· | ···················· | ···················· | ···················· |
| AB | 6172 | hCoV-19/Netherlands/ZH-EMC-1172/2020\|EPI_ISL_76324 | ···················· | ···················· | ···················· | ···················· | ···················· | ···················· |
| AB | 6173 | hCoV-19/Netherlands/ZH-EMC-1173/2020\|EPI_ISL_76324 | ···················· | ···················· | ···················· | ···················· | ···················· | ···················· |
| AB | 6174 | hCoV-19/Netherlands/ZH-EMC-1175/2020\|EPI_ISL_76325 | ···················· | ···················· | ···················· | ···················· | ···················· | ···················· |
| AB | 6175 | hCoV-19/Netherlands/ZH-EMC-1177/2020\|EPI_ISL_76325 | ···················· | ···················· | ···················· | ···················· | ···················· | ···················· |
| AB | 6176 | hCoV-19/Netherlands/ZH-EMC-1179/2020\|EPI_ISL_76325 | ···················· | ···················· | ···················· | ···················· | ···················· | ···················· |
| AB | 6177 | hCoV-19/Netherlands/ZH-EMC-1180/2020\|EPI_ISL_76325 | ···················· | ···················· | ···················· | ···················· | ···················· | ···················· |
| AB | 6178 | hCoV-19/Netherlands/ZH-EMC-1182/2020\|EPI_ISL_76325 | ···················· | ···················· | ···················· | ···················· | ···················· | ···················· |
| AB | 6179 | hCoV-19/Netherlands/ZH-EMC-1183/2020\|EPI_ISL_76325 | ···················· | ···················· | ···················· | ···················· | ···················· | ···················· |
| AB | 6180 | hCoV-19/England/NOTT-121828/2020\|EPI_ISL_764502\|20 | ···················· | ·A·······A·········· | ···················· | ···················· | ···················· | ···················· |
| AB | 6181 | hCoV-19/England/NOTT-121830/2020\|EPI_ISL_764503\|20 | ···················· | ···················· | ···················· | ···················· | ···················· | ···················· |
| AB | 6182 | hCoV-19/England/NOTT-121833/2020\|EPI_ISL_764504\|20 | ···················· | ···················· | ···················· | ···················· | ···················· | ···················· |
| AB | 6183 | hCoV-19/England/NOTT-121835/2020\|EPI_ISL_764505\|20 | ···················· | ···················· | ···················· | ···················· | ···················· | ···················· |
| AB | 6184 | hCoV-19/England/NOTT-121836/2020\|EPI_ISL_764506\|20 | ···················· | ···················· | ···················· | ···················· | ···················· | ···················· |
| AB | 6185 | hCoV-19/England/NOTT-121837/2020\|EPI_ISL_764507\|20 | ···················· | ···················· | ···················· | ···················· | ···················· | ···················· |
| AB | 6186 | hCoV-19/England/NOTT-121840/2020\|EPI_ISL_764508\|20 | ···················· | ···················· | ···················· | ···················· | ···················· | ···················· |
| AB | 6187 | hCoV-19/England/NOTT-121842/2020\|EPI_ISL_764509\|20 | ···················· | ···················· | ···················· | ···················· | ···················· | ···················· |
| AB | 6188 | hCoV-19/England/NOTT-121846/2020\|EPI_ISL_764510\|20 | ···················· | ···················· | ···················· | ···················· | ···················· | ···················· |
| AB | 6189 | hCoV-19/England/NOTT-121850/2020\|EPI_ISL_764511\|20 | ···················· | ···················· | ···················· | ···················· | ···················· | ···················· |
| AB | 6190 | hCoV-19/England/NOTT-121851/2020\|EPI_ISL_764512\|20 | ···················· | ···················· | ···················· | ···················· | ···················· | ···················· |
| AB | 6191 | hCoV-19/England/NOTT-121856/2020\|EPI_ISL_764513\|20 | ···················· | ···················· | ···················· | ···················· | ···················· | ···················· |
| AB | 6192 | hCoV-19/England/NOTT-121857/2020\|EPI_ISL_764514\|20 | ···················· | ···················· | ···················· | ···················· | ···················· | ···················· |
| AB | 6193 | hCoV-19/England/NOTT-121859/2020\|EPI_ISL_764515\|20 | ···················· | ···················· | ···················· | ···················· | ···················· | ···················· |
| AB | 6194 | hCoV-19/England/NOTT-121861/2020\|EPI_ISL_764516\|20 | ···················· | ···················· | ···················· | ···················· | ···················· | ···················· |
| AB | 6195 | hCoV-19/England/NOTT-121862/2020\|EPI_ISL_764517\|20 | ···················· | ···················· | ···················· | ···················· | ···················· | ···················· |
| AB | 6196 | hCoV-19/England/NOTT-121865/2020\|EPI_ISL_764518\|20 | ···················· | ···················· | ···················· | ···················· | ···················· | ···················· |
| AB | 6197 | hCoV-19/England/NOTT-121866/2020\|EPI_ISL_764519\|20 | ···················· | ···················· | ···················· | ···················· | ···················· | ···················· |
| AB | 6198 | hCoV-19/England/NOTT-121868/2020\|EPI_ISL_764520\|20 | ···················· | ···················· | ···················· | ···················· | ···················· | ···················· |
| AB | 6199 | hCoV-19/England/NOTT-121869/2020\|EPI_ISL_764521\|20 | ···················· | ···················· | ···················· | ···················· | ···················· | ···················· |
| AB | 6200 | hCoV-19/England/NOTT-121936/2020\|EPI_ISL_764522\|20 | ···················· | ···················· | ···················· | ···················· | ···················· | ···················· |
| AB | 6201 | hCoV-19/England/NOTT-121949/2020\|EPI_ISL_764523\|20 | ···················· | ···················· | ···················· | ···················· | ···················· | ···················· |
| AB | 6202 | hCoV-19/England/NOTT-121952/2020\|EPI_ISL_764524\|20 | ···················· | ···················· | ···················· | ···················· | ···················· | ···················· |
| AB | 6203 | hCoV-19/England/OXON-F3FF39/2020\|EPI_ISL_764525\|20 | ···················· | ···················· | ···················· | ···················· | ···················· | ···················· |
| AB | 6204 | hCoV-19/England/OXON-F40032/2020\|EPI_ISL_764526\|20 | ···················· | ···················· | ···················· | ···················· | ···················· | ···················· |
| AB | 6205 | hCoV-19/England/OXON-F40041/2020\|EPI_ISL_764527\|20 | ···················· | ···················· | ···················· | ···················· | ···················· | ···················· |
| AB | 6206 | hCoV-19/England/OXON-F4007E/2020\|EPI_ISL_764528\|20 | ···················· | ···················· | ···················· | ···················· | ···················· | ···················· |
| AB | 6207 | hCoV-19/England/OXON-F4009C/2020\|EPI_ISL_764529\|20 | ···················· | ···················· | ···················· | ···················· | ···················· | ···················· |
| AB | 6208 | hCoV-19/Scotland/OXON-F401F3/2020\|EPI_ISL_764530\|2 | ···················· | ···················· | ···················· | ···················· | ···················· | ···················· |
| AB | 6209 | hCoV-19/Scotland/OXON-F4024B/2020\|EPI_ISL_764531\|2 | ···················· | ···················· | ···················· | ···················· | ···················· | ···················· |
| AB | 6210 | hCoV-19/Northern Ireland/OXON-F40533/2020\|EPI_ISL_ | ···················· | ···················· | ···················· | ···················· | ···················· | ···················· |
| AB | 6211 | hCoV-19/England/OXON-F4067C/2020\|EPI_ISL_764533\|20 | ···················· | ···················· | ···················· | ···················· | ···················· | ···················· |
| AB | 6212 | hCoV-19/Wales/OXON-F4075B/2020\|EPI_ISL_764534\|2020 | ···················· | ···················· | ···················· | ···················· | ···················· | ···················· |
| AB | 6213 | hCoV-19/England/OXON-F40BC8/2020\|EPI_ISL_764546\|20 | ···················· | ···················· | ···················· | ···················· | ···················· | ···················· |
| AB | 6214 | hCoV-19/England/OXON-F40BF5/2020\|EPI_ISL_764547\|20 | ···················· | ···················· | ···················· | ···················· | ······T············· | ···················· |
| AB | 6215 | hCoV-19/England/OXON-F40C2F/2020\|EPI_ISL_764548\|20 | ···················· | ···················· | ···················· | ···················· | ···················· | ···················· |
| AB | 6216 | hCoV-19/England/OXON-F40CA7/2020\|EPI_ISL_764549\|20 | ···················· | ···················· | ···················· | ···················· | ···················· | ···················· |
| AB | 6217 | hCoV-19/Wales/OXON-F40CB6/2020\|EPI_ISL_764550\|2020 | ···················· | ···················· | ···················· | ···················· | ···················· | ···················· |
| AB | 6218 | hCoV-19/England/OXON-F40D77/2020\|EPI_ISL_764551\|20 | ···················· | ···················· | ···················· | ···················· | ···················· | ···················· |
| AB | 6219 | hCoV-19/Wales/OXON-F40DC2/2020\|EPI_ISL_764552\|2020 | ···················· | ···················· | ···················· | ···················· | ···················· | ···················· |
| AB | 6220 | hCoV-19/Wales/OXON-F40E65/2020\|EPI_ISL_764553\|2020 | ···················· | ···················· | ···················· | ···················· | ···················· | ···················· |
| AB | 6221 | hCoV-19/England/OXON-F40EFC/2020\|EPI_ISL_764554\|20 | ···················· | ···················· | ···················· | ···················· | ···················· | ···················· |
| AB | 6222 | hCoV-19/England/OXON-F40F44/2020\|EPI_ISL_764556\|20 | ···················· | ···················· | ···················· | ···················· | ···················· | ···················· |
| AB | 6223 | hCoV-19/England/OXON-F40F53/2020\|EPI_ISL_764557\|20 | ···················· | ···················· | ···················· | ···················· | ···················· | ···················· |
| AB | 6224 | hCoV-19/England/OXON-F410C8/2020\|EPI_ISL_764558\|20 | ···················· | ···················· | ···················· | ···················· | ···················· | ···················· |
| AB | 6225 | hCoV-19/England/OXON-F410D7/2020\|EPI_ISL_764559\|20 | ···················· | ···················· | ···················· | ···················· | ···················· | ···················· |

| | | | | | | | | |
|---|---|---|---|---|---|---|---|---|
| AB | 6226 | hCoV-19/England/OXON-F411D4/2020\|EPI_ISL_764560\|20 | .................... | .................... | .................... | .................... | .................... | .................... |
| AB | 6227 | hCoV-19/England/OXON-F4122C/2020\|EPI_ISL_764561\|20 | .................... | .................... | .................... | .................... | .................... | .................... |
| AB | 6228 | hCoV-19/England/OXON-F41268/2020\|EPI_ISL_764562\|20 | .................... | .................... | .................... | .................... | .................... | .................... |
| AB | 6229 | hCoV-19/England/OXON-F41295/2020\|EPI_ISL_764563\|20 | .................... | .................... | .................... | .................... | .................... | .................... |
| AB | 6230 | hCoV-19/England/OXON-F412D1/2020\|EPI_ISL_764564\|20 | .................... | .................... | .................... | .................... | .................... | .................... |
| AB | 6231 | hCoV-19/England/OXON-F41444/2020\|EPI_ISL_764565\|20 | .................... | .................... | .................... | .................... | .................... | .................... |
| AB | 6232 | hCoV-19/Scotland/OXON-F41480/2020\|EPI_ISL_764566\|2 | .................... | .................... | .................... | .................... | .................... | .................... |
| AB | 6233 | hCoV-19/England/OXON-F4149F/2020\|EPI_ISL_764567\|20 | .................... | .................... | .................... | .................... | .................... | .................... |
| AB | 6234 | hCoV-19/England/OXON-F414CC/2020\|EPI_ISL_764568\|20 | .................... | .................... | .................... | .................... | .................... | .................... |
| AB | 6235 | hCoV-19/England/OXON-F414DB/2020\|EPI_ISL_764569\|20 | .................... | .................... | .................... | .................... | .................... | .................... |
| AB | 6236 | hCoV-19/England/OXON-F414EA/2020\|EPI_ISL_764570\|20 | .................... | .................... | .................... | .................... | .................... | .................... |
| AB | 6237 | hCoV-19/Scotland/OXON-F4157E/2020\|EPI_ISL_764571\|2 | .................... | .................... | .................... | .................... | .................... | .................... |
| AB | 6238 | hCoV-19/England/OXON-F415E7/2020\|EPI_ISL_764572\|20 | .................... | .................... | .................... | .................... | .................... | .................... |
| AB | 6239 | hCoV-19/Wales/PHWC-4A379D/2020\|EPI_ISL_764573\|2020 | .................... | .................... | .................... | .................... | .................... | .................... |
| AB | 6240 | hCoV-19/Wales/PHWC-4A37BB/2020\|EPI_ISL_764574\|2020 | .................... | .................... | .................... | .................... | .................... | .................... |
| AB | 6241 | hCoV-19/Wales/PHWC-4A37CA/2020\|EPI_ISL_764575\|2020 | .................... | .................... | .................... | .................... | .................... | .................... |
| AB | 6242 | hCoV-19/Wales/PHWC-4A37E8/2020\|EPI_ISL_764576\|2020 | .................... | .................... | .................... | .................... | .................... | .................... |
| AB | 6243 | hCoV-19/Wales/PHWC-4A37F7/2020\|EPI_ISL_764577\|2020 | .................... | .................... | .................... | .................... | .................... | .................... |
| AB | 6244 | hCoV-19/Wales/PHWC-4A3803/2020\|EPI_ISL_764578\|2020 | .................... | .................... | .................... | .................... | .................... | .................... |
| AB | 6245 | hCoV-19/Wales/PHWC-4A3812/2020\|EPI_ISL_764579\|2020 | .................... | .................... | .................... | .................... | .................... | .................... |
| AB | 6246 | hCoV-19/Wales/PHWC-4A3821/2020\|EPI_ISL_764580\|2020 | .................... | .................... | .................... | .................... | .................... | .................... |
| AB | 6247 | hCoV-19/Wales/PHWC-4A3830/2020\|EPI_ISL_764581\|2020 | .................... | .................... | .................... | .................... | .................... | .................... |
| AB | 6250 | hCoV-19/Wales/PHWC-4A388B/2020\|EPI_ISL_764584\|2020 | .................... | .................... | .................... | .................... | .................... | .................... |
| AB | 6251 | hCoV-19/Wales/PHWC-4A38A9/2020\|EPI_ISL_764585\|2020 | .................... | .................... | .................... | .................... | .................... | .................... |
| AB | 6252 | hCoV-19/Wales/PHWC-4A38C7/2020\|EPI_ISL_764586\|2020 | .................... | .................... | .................... | .................... | .................... | .................... |
| AB | 6255 | hCoV-19/Wales/PHWC-4A3900/2020\|EPI_ISL_764589\|2020 | .................... | .................... | .................... | .................... | .................... | .................... |
| AB | 6256 | hCoV-19/USA/MA-MGH-03428/2020\|EPI_ISL_765833\|2020- | .................... | .................... | .................... | .................... | .................... | .................... |
| AB | 6257 | hCoV-19/USA/MA-MGH-03429/2020\|EPI_ISL_765834\|2020- | .................... | .A.................. | .................... | .................... | .................... | .................... |
| AB | 6258 | hCoV-19/USA/MA-MGH-03431/2020\|EPI_ISL_765835\|2020- | .................... | .................... | .................... | .................... | .................... | .................... |
| AB | 6259 | hCoV-19/USA/MA-MGH-03434/2020\|EPI_ISL_765836\|2020- | .................... | .................... | .................... | .................... | .................... | .................... |
| AB | 6260 | hCoV-19/USA/MA-MGH-03435/2020\|EPI_ISL_765837\|2020- | .................... | .................... | .................... | .................... | .................... | .................... |
| AB | 6261 | hCoV-19/USA/MA-MGH-03437/2020\|EPI_ISL_765838\|2020- | .................... | .................... | .................... | .................... | .................... | .................... |
| AB | 6262 | hCoV-19/USA/MA-MGH-03442/2020\|EPI_ISL_765839\|2020- | .................... | .................... | .................... | .................... | .................... | .................... |
| AB | 6263 | hCoV-19/USA/MA-MGH-03445/2020\|EPI_ISL_765840\|2020- | .................... | .................... | .................... | .................... | .................... | .................... |
| AB | 6264 | hCoV-19/USA/MA-MGH-03450/2020\|EPI_ISL_765842\|2020- | .................... | .................... | .................... | .................... | .................... | .................... |
| AB | 6265 | hCoV-19/USA/MA-MGH-03451/2020\|EPI_ISL_765843\|2020- | .................... | .A.................. | .................... | .................... | .................... | .................... |
| AB | 6266 | hCoV-19/USA/MA-MGH-03452/2020\|EPI_ISL_765844\|2020- | .................... | .................... | .................... | .................... | .................... | .................... |
| AB | 6267 | hCoV-19/USA/MA-MGH-03460/2020\|EPI_ISL_765845\|2020- | .................... | .................... | .................... | .................... | .................... | .................... |
| AB | 6268 | hCoV-19/USA/MA-MGH-03462/2020\|EPI_ISL_765846\|2020- | .................... | .................... | .................... | .................... | .................... | .................... |
| AB | 6269 | hCoV-19/USA/MA-MGH-03464/2020\|EPI_ISL_765847\|2020- | .................... | .................... | .................... | .................... | .................... | .................... |
| AB | 6270 | hCoV-19/USA/MA-MGH-03465/2020\|EPI_ISL_765848\|2020- | .................... | .................... | .................... | .................... | .................... | .................... |
| AB | 6271 | hCoV-19/USA/MA-MGH-03466/2020\|EPI_ISL_765849\|2020- | .................... | .................... | .................... | .................... | .................... | .................... |
| AB | 6272 | hCoV-19/USA/MA-MGH-03468/2020\|EPI_ISL_765850\|2020- | .................... | .A.................. | .................... | .................... | .................... | .................... |
| AB | 6273 | hCoV-19/USA/MA-MGH-03470/2020\|EPI_ISL_765851\|2020- | .................... | .................... | .................... | .................... | .................... | .................... |
| AB | 6274 | hCoV-19/USA/MA-MGH-03471/2020\|EPI_ISL_765852\|2020- | .................... | .................... | .................... | .................... | .................... | .................... |
| AB | 6275 | hCoV-19/USA/MA-MGH-03475/2020\|EPI_ISL_765853\|2020- | .................... | .A.................. | .................... | .................... | .................... | .................... |
| AB | 6276 | hCoV-19/USA/MA-MGH-03476/2020\|EPI_ISL_765854\|2020- | .................... | .A.................. | .................... | .................... | .................... | .................... |
| AB | 6277 | hCoV-19/USA/MA-MGH-03479/2020\|EPI_ISL_765855\|2020- | .................... | .................... | .................... | .................... | .................... | .................... |
| AB | 6278 | hCoV-19/USA/MA-MGH-03483/2020\|EPI_ISL_765856\|2020- | .................... | .................... | .................... | .................... | .................... | .................... |
| AB | 6279 | hCoV-19/USA/MA-MGH-03485/2020\|EPI_ISL_765857\|2020- | .................... | .................... | .................... | .................... | .................... | .................... |
| AB | 6280 | hCoV-19/USA/MA-MGH-03486/2020\|EPI_ISL_765858\|2020- | .................... | .............A...... | .................... | .................... | .................... | .................... |
| AB | 6281 | hCoV-19/USA/MA-MGH-03494/2020\|EPI_ISL_765860\|2020- | .................... | .................... | .................... | .................... | .................... | .................... |
| AB | 6282 | hCoV-19/USA/MA-MGH-03495/2020\|EPI_ISL_765861\|2020- | .................... | .A.................. | .................... | .................... | .................... | .................... |
| AB | 6283 | hCoV-19/USA/MA-MGH-03496/2020\|EPI_ISL_765862\|2020- | .................... | .................... | .................... | .................... | .................... | .................... |
| AB | 6284 | hCoV-19/USA/MA-MGH-03503/2020\|EPI_ISL_765863\|2020- | .................... | .................... | .................... | .................... | .................... | .................... |
| AB | 6285 | hCoV-19/USA/MA-MGH-03504/2020\|EPI_ISL_765864\|2020- | .................... | .................... | .................... | .................... | .................... | .................... |
| AB | 6286 | hCoV-19/USA/MA-MGH-03506/2020\|EPI_ISL_765865\|2020- | .................... | .................... | .................... | .................... | .................... | .................... |
| AB | 6287 | hCoV-19/USA/MA-MGH-03531/2020\|EPI_ISL_765875\|2020- | .................... | .................... | .................... | .................... | .................... | .................... |
| AB | 6288 | hCoV-19/USA/MA-MGH-03537/2020\|EPI_ISL_765877\|2020- | .................... | .................... | .................... | .................... | .................... | .................... |
| AB | 6289 | hCoV-19/USA/MA-MGH-03542/2020\|EPI_ISL_765878\|2020- | .................... | .................... | .................... | .................... | .................... | .................... |
| AB | 6290 | hCoV-19/USA/MA-MGH-03543/2020\|EPI_ISL_765879\|2020- | .................... | .................... | .................... | .................... | .................... | .................... |
| AB | 6291 | hCoV-19/USA/MA-MGH-03544/2020\|EPI_ISL_765880\|2020- | .................... | .A.................. | .................... | .................... | .................... | .................... |
| AB | 6292 | hCoV-19/USA/MA-MGH-03553/2020\|EPI_ISL_765881\|2020- | .................... | .................... | .................... | .................... | .................... | .................... |
| AB | 6293 | hCoV-19/USA/MA-MGH-03555/2020\|EPI_ISL_765882\|2020- | .................... | .................... | .................... | .................... | .................... | .................... |
| AB | 6294 | hCoV-19/USA/MA-MGH-03558/2020\|EPI_ISL_765883\|2020- | .................... | .................... | .................... | .................... | .................... | .................... |
| AB | 6295 | hCoV-19/USA/MA-MGH-03561/2020\|EPI_ISL_765884\|2020- | .................... | .................... | .................... | .................... | .................... | .................... |
| AB | 6296 | hCoV-19/USA/MA-MGH-03563/2020\|EPI_ISL_765885\|2020- | .................... | .................... | .................... | .................... | .................... | .................... |
| AB | 6297 | hCoV-19/USA/MA-MGH-03565/2020\|EPI_ISL_765887\|2020- | .................... | .................... | .................... | .................... | .................... | .................... |
| AB | 6298 | hCoV-19/USA/MA-MGH-03570/2020\|EPI_ISL_765889\|2020- | .................... | .................... | .................... | .................... | .................... | .................... |
| AB | 6299 | hCoV-19/USA/MA-MGH-03583/2020\|EPI_ISL_765892\|2020- | .................... | .................... | .................... | .................... | .................... | .................... |
| AB | 6300 | hCoV-19/Canada/ON-PHL-20-03719/2020\|EPI_ISL_775200 | .................... | .................... | .................... | .................... | .................... | .................... |
| AB | 6301 | hCoV-19/Canada/ON-PHL-20-03720/2020\|EPI_ISL_775201 | .................... | .................... | .................... | .................... | .................... | .................... |
| AB | 6302 | hCoV-19/Canada/ON-PHL-20-03721/2020\|EPI_ISL_775202 | .................... | .................... | .................... | .................... | .................... | .................... |
| AB | 6303 | hCoV-19/Canada/ON-PHL-20-03723/2020\|EPI_ISL_775203 | .................... | .................... | .................... | .................... | .................... | .................... |
| AB | 6304 | hCoV-19/Canada/ON-PHL-20-03724/2020\|EPI_ISL_775204 | .................... | .................... | .................... | .................... | .................... | .................... |
| AB | 6305 | hCoV-19/Canada/ON-PHL-20-03742/2020\|EPI_ISL_775210 | .................... | .................... | .................... | .................... | .................... | .................... |
| AB | 6306 | hCoV-19/Canada/ON-PHL-20-03743/2020\|EPI_ISL_775211 | .................... | .................... | .................... | .................... | .................... | .................... |
| AB | 6307 | hCoV-19/Canada/ON-PHL-20-03744/2020\|EPI_ISL_775212 | .................... | .................... | .................... | .................... | .................... | .................... |
| AB | 6308 | hCoV-19/Bangladesh/GRMDRC-4/2020\|EPI_ISL_775213\|20 | .................... | .................... | .................... | .................... | .................... | .................... |
| AB | 6309 | hCoV-19/Bangladesh/GRMDRC-6/2020\|EPI_ISL_775215\|20 | .................... | .................... | .................... | .................... | .................... | .................... |

| | | | | | | | | |
|---|---|---|---|---|---|---|---|---|
| AB | 6310 | hCoV-19/Bangladesh/GRMDRC-7/2020\|EPI_ISL_775216\|20 | ···················· | ···················· | ···················· | ···················· | ···················· | ···················· |
| AB | 6311 | hCoV-19/Bangladesh/GRMDRC-8/2020\|EPI_ISL_775217\|20 | ···················· | ···················· | ···················· | ···················· | ···················· | ···················· |
| AB | 6312 | hCoV-19/Bangladesh/GRMDRC-9/2020\|EPI_ISL_775218\|20 | ···················· | ···················· | ···················· | ···················· | ···················· | ···················· |
| AB | 6313 | hCoV-19/Morocco/FMP-44/2020\|EPI_ISL_775220\|2020-12 | ···················· | ···················· | ···················· | ···················· | ···················· | ···················· |
| AB | 6314 | hCoV-19/Morocco/FMP-45/2020\|EPI_ISL_775221\|2020-12 | ···················· | ···················· | ···················· | ···················· | ···················· | ···················· |
| AB | 6315 | hCoV-19/Morocco/FMP-46/2020\|EPI_ISL_775222\|2020-12 | ···················· | ···················· | ···················· | ···················· | ···················· | ···················· |
| AB | 6316 | hCoV-19/Morocco/FMP-47/2020\|EPI_ISL_775223\|2020-12 | ···················· | ···················· | ···················· | ···················· | ···················· | ···················· |
| AB | 6317 | hCoV-19/Morocco/FMP-48/2020\|EPI_ISL_775224\|2020-12 | ···················· | ···················· | ···················· | ···················· | ···················· | ···················· |
| AB | 6318 | hCoV-19/Morocco/FMP-49/2020\|EPI_ISL_775225\|2020-12 | ···················· | ···················· | ···················· | ···················· | ···················· | ···················· |
| AB | 6319 | hCoV-19/Morocco/FMP-50/2020\|EPI_ISL_775226\|2020-12 | ···················· | ···················· | ···················· | ···················· | ···················· | ···················· |
| AB | 6320 | hCoV-19/Germany/HH-hpi-p2431/2020\|EPI_ISL_776500\|2 | ···················· | ···················· | ···················· | ···················· | ···················· | ···················· |
| AB | 6322 | hCoV-19/Germany/HH-hpi-p2435/2020\|EPI_ISL_776502\|2 | ···················· | ···················· | ···················· | ···················· | ···················· | ···················· |
| AB | 6323 | hCoV-19/Germany/HH-hpi-p2436/2020\|EPI_ISL_776503\|2 | ···················· | ···················· | ···················· | ···················· | ···················· | ···················· |
| AB | 6324 | hCoV-19/Germany/HH-hpi-p2437/2020\|EPI_ISL_776504\|2 | ···················· | ···················· | ···················· | ···················· | ···················· | ···················· |
| AB | 6325 | hCoV-19/Germany/HH-hpi-p2439/2020\|EPI_ISL_776505\|2 | ···················· | ···················· | ···················· | ···················· | ···················· | ···················· |
| AB | 6327 | hCoV-19/Germany/HH-hpi-p2441/2020\|EPI_ISL_776507\|2 | ···················· | ···················· | ···················· | ···················· | ···················· | ···················· |
| AB | 6328 | hCoV-19/Germany/HH-hpi-p2443/2020\|EPI_ISL_776508\|2 | ···················· | ···················· | ···················· | ···················· | ···················· | ···················· |
| AB | 6329 | hCoV-19/Germany/HH-hpi-p2444/2020\|EPI_ISL_776509\|2 | ···················· | ···················· | ···················· | ···················· | ···················· | ···················· |
| AB | 6330 | hCoV-19/Germany/HH-hpi-p2452/2020\|EPI_ISL_776514\|2 | ···················· | ···················· | ···················· | ···················· | ···················· | ···················· |
| AB | 6331 | hCoV-19/Germany/HH-hpi-p2453/2020\|EPI_ISL_776515\|2 | ···················· | ···················· | ···················· | ···················· | ···················· | ···················· |
| AB | 6332 | hCoV-19/Germany/HH-hpi-p2454/2020\|EPI_ISL_776516\|2 | ···················· | ···················· | ···················· | ···················· | ···················· | ···················· |
| AB | 6333 | hCoV-19/Germany/HH-hpi-p2455/2020\|EPI_ISL_776517\|2 | ···················· | ···················· | ···················· | ···················· | ···················· | ···················· |
| AB | 6334 | hCoV-19/Germany/HH-hpi-p2457/2020\|EPI_ISL_776518\|2 | ···················· | ···················· | ···················· | ···················· | ···················· | ···················· |
| AB | 6335 | hCoV-19/Germany/HH-hpi-p2458/2020\|EPI_ISL_776519\|2 | ···················· | ···················· | ···················· | ···················· | ···················· | ···················· |
| AB | 6337 | hCoV-19/Germany/HH-hpi-p2462/2020\|EPI_ISL_776521\|2 | ···················· | ···················· | ···················· | ···················· | ···················· | ···················· |
| AB | 6339 | hCoV-19/Germany/HH-hpi-p2464/2020\|EPI_ISL_776523\|2 | ···················· | ···················· | ···················· | ···················· | ···················· | ···················· |
| AB | 6340 | hCoV-19/Germany/HH-hpi-p2465/2020\|EPI_ISL_776524\|2 | ···················· | ···················· | ···················· | ···················· | ···················· | ···················· |
| AB | 6341 | hCoV-19/Germany/HH-hpi-p2466/2020\|EPI_ISL_776525\|2 | ···················· | ···················· | ···················· | ···················· | ···················· | ···················· |
| AB | 6342 | hCoV-19/Germany/HH-hpi-p2467/2020\|EPI_ISL_776526\|2 | ···················· | ···················· | ···················· | ···················· | ···················· | ···················· |
| AB | 6343 | hCoV-19/Germany/HH-hpi-p2468/2020\|EPI_ISL_776527\|2 | ···················· | ···················· | ···················· | ···················· | ···················· | ···················· |
| AB | 6344 | hCoV-19/Germany/HH-hpi-p2469/2020\|EPI_ISL_776528\|2 | ···················· | ···················· | ···················· | ···················· | ···················· | ···················· |
| AB | 6345 | hCoV-19/Germany/HH-hpi-p2471/2020\|EPI_ISL_776529\|2 | ···················· | ···················· | ···················· | ···················· | ···················· | ···················· |
| AB | 6346 | hCoV-19/Germany/HH-hpi-p2472/2020\|EPI_ISL_776530\|2 | ···················· | ···················· | ···················· | ···················· | ···················· | ···················· |
| AB | 6347 | hCoV-19/Germany/HH-hpi-p2473/2020\|EPI_ISL_776531\|2 | ···················· | ···················· | ···················· | ···················· | ···················· | ···················· |
| AB | 6348 | hCoV-19/Germany/HH-hpi-p2474/2020\|EPI_ISL_776532\|2 | ···················· | ···················· | ···················· | ···················· | ···················· | ···················· |
| AB | 6349 | hCoV-19/Germany/HH-hpi-p2475/2020\|EPI_ISL_776533\|2 | ···················· | ···················· | ···················· | ···················· | ···················· | ···················· |
| AB | 6350 | hCoV-19/Germany/HH-hpi-p2476/2020\|EPI_ISL_776534\|2 | ···················· | ···················· | ···················· | ···················· | ···················· | ···················· |
| AB | 6351 | hCoV-19/Germany/HH-hpi-p2477/2020\|EPI_ISL_776535\|2 | ···················· | ···················· | ···················· | ···················· | ···················· | ···················· |
| AB | 6352 | hCoV-19/Germany/HH-hpi-p2483/2020\|EPI_ISL_776540\|2 | ···················· | ···················· | ···················· | ···················· | ···················· | ···················· |
| AB | 6353 | hCoV-19/Germany/HH-hpi-p2484/2020\|EPI_ISL_776541\|2 | ···················· | ···················· | ···················· | ···················· | ···················· | ···················· |
| AB | 6354 | hCoV-19/Germany/HH-hpi-p2485/2020\|EPI_ISL_776542\|2 | ···················· | ···················· | ···················· | ···················· | ···················· | ···················· |
| AB | 6356 | hCoV-19/Germany/HH-hpi-p2494/2020\|EPI_ISL_776544\|2 | ···················· | ···················· | ···················· | ···················· | ···················· | ···················· |
| AB | 6357 | hCoV-19/Germany/HH-hpi-p2495/2020\|EPI_ISL_776545\|2 | ···················· | ···················· | ···················· | ···················· | ···················· | ···················· |
| AB | 6358 | hCoV-19/Germany/HH-hpi-p2496/2020\|EPI_ISL_776546\|2 | ···················· | ···················· | ···················· | ···················· | ···················· | ···················· |
| AB | 6359 | hCoV-19/Germany/HH-hpi-p2497/2020\|EPI_ISL_776547\|2 | ···················· | ···················· | ···················· | ···················· | ···················· | ···················· |
| AB | 6360 | hCoV-19/Germany/NI-hpi-p1458/2020\|EPI_ISL_776548\|2 | ···················· | ···················· | ···················· | ···················· | ···················· | ···················· |
| AB | 6361 | hCoV-19/Germany/NI-hpi-p1552/2020\|EPI_ISL_776549\|2 | ···················· | ···················· | ···················· | ···················· | ···················· | ···················· |
| AB | 6362 | hCoV-19/Germany/NI-hpi-p1674/2020\|EPI_ISL_776550\|2 | ···················· | ···················· | ···················· | ···················· | ···················· | ···················· |
| AB | 6363 | hCoV-19/Germany/NI-hpi-p1718/2020\|EPI_ISL_776551\|2 | ···················· | ···················· | ···················· | ···················· | ···················· | ···················· |
| AB | 6364 | hCoV-19/Germany/NI-hpi-p1746/2020\|EPI_ISL_776552\|2 | ···················· | ···················· | ···················· | ···················· | ···················· | ···················· |
| AB | 6366 | hCoV-19/Germany/RP-hpi-p2411/2020\|EPI_ISL_776554\|2 | ···················· | ···················· | ···················· | ···················· | ···················· | ···················· |
| AB | 6367 | hCoV-19/Germany/SH-hpi-p1578/2020\|EPI_ISL_776555\|2 | ···················· | ···················· | ···················· | ···················· | ···················· | ···················· |
| AB | 6368 | hCoV-19/Germany/SH-hpi-p1886/2020\|EPI_ISL_776556\|2 | ···················· | ···················· | ···················· | ···················· | ···················· | ···················· |
| AB | 6369 | hCoV-19/Germany/SH-hpi-p2017/2020\|EPI_ISL_776557\|2 | ···················· | ···················· | ···················· | ···················· | ···················· | ···················· |
| AB | 6370 | hCoV-19/Germany/SH-hpi-p2460/2020\|EPI_ISL_776558\|2 | ···················· | ···················· | ···················· | ···················· | ···················· | ···················· |
| AB | 6371 | hCoV-19/Germany/TH-hpi-p1690/2020\|EPI_ISL_776559\|2 | ···················· | ···················· | ···················· | ···················· | ···················· | ···················· |
| AB | 6372 | hCoV-19/Germany/HH-hpi-p1364/2020\|EPI_ISL_776560\|2 | ···················· | ···················· | ···················· | ···················· | ···················· | ···················· |
| AB | 6373 | hCoV-19/Germany/HH-hpi-p1699/2020\|EPI_ISL_776561\|2 | ···················· | ···················· | ···················· | ···················· | ···················· | ···················· |
| AB | 6374 | hCoV-19/Germany/HH-hpi-p2314/2020\|EPI_ISL_776562\|2 | ···················· | ···················· | ···················· | ···················· | ···················· | ···················· |
| AB | 6375 | hCoV-19/Germany/HH-hpi-p2317/2020\|EPI_ISL_776563\|2 | ···················· | ···················· | ···················· | ···················· | ···················· | ···················· |
| AB | 6376 | hCoV-19/Germany/HH-hpi-p1939/2020\|EPI_ISL_776564\|2 | ···················· | ···················· | ···················· | ···················· | ···················· | ···················· |
| AB | 6377 | hCoV-19/Germany/SH-hpi-p1907/2020\|EPI_ISL_776565\|2 | ···················· | ···················· | ···················· | ···················· | ···················· | ···················· |
| AB | 6378 | hCoV-19/Germany/SH-hpi-p2305/2020\|EPI_ISL_776566\|2 | ···················· | ···················· | ···················· | ···················· | ···················· | ···················· |
| AB | 6379 | hCoV-19/USA/OH-ODH-SC35093/2020\|EPI_ISL_776568\|202 | ···················· | ···················· | ···················· | ···················· | ···················· | ···················· |
| AB | 6380 | hCoV-19/England/CAMC-CB9FC1/2020\|EPI_ISL_777814\|20 | ···················· | ···················· | ···················· | ···················· | ···················· | ···················· |
| AB | 6381 | hCoV-19/England/CAMC-CB89B5/2020\|EPI_ISL_777819\|20 | ···················· | ···················· | ···················· | ···················· | ······T············· | ···················· |
| AB | 6382 | hCoV-19/England/CAMC-CB8812/2020\|EPI_ISL_777820\|20 | ···················· | ···················· | ···················· | ···················· | ···················· | ···················· |
| AB | 6383 | hCoV-19/England/CAMC-CB6B57/2020\|EPI_ISL_777824\|20 | ···················· | ·A·················· | ···················· | ···················· | ···················· | ···················· |
| AB | 6384 | hCoV-19/England/CAMC-CB786E/2020\|EPI_ISL_777829\|20 | ···················· | ···················· | ···················· | ···················· | ···················· | ···················· |
| AB | 6385 | hCoV-19/England/CAMC-CB9ED3/2020\|EPI_ISL_777832\|20 | ···················· | ···················· | ···················· | ···················· | ···················· | ···················· |
| AB | 6386 | hCoV-19/England/CAMC-CB8C52/2020\|EPI_ISL_777836\|20 | ···················· | ···················· | ···················· | ···················· | ···················· | ···················· |
| AB | 6387 | hCoV-19/England/CAMC-CB892E/2020\|EPI_ISL_777845\|20 | ···················· | ···················· | ···················· | ···················· | ···················· | ···················· |
| AB | 6388 | hCoV-19/England/CAMC-CB7716/2020\|EPI_ISL_777847\|20 | ···················· | ·A·················· | ···················· | ···················· | ···················· | ···················· |
| AB | 6389 | hCoV-19/England/CAMC-CB8AB2/2020\|EPI_ISL_777848\|20 | ···················· | ···················· | ···················· | ···················· | ···················· | ···················· |
| AB | 6390 | hCoV-19/England/CAMC-CB7D14/2020\|EPI_ISL_777852\|20 | ···················· | ···················· | ···················· | ···················· | ···················· | ···················· |
| AB | 6391 | hCoV-19/England/CAMC-CBA054/2020\|EPI_ISL_777856\|20 | ···················· | ···················· | ···················· | ···················· | ···················· | ···················· |
| AB | 6392 | hCoV-19/England/CAMC-CBA036/2020\|EPI_ISL_777857\|20 | ···················· | ···················· | ···················· | ···················· | ···················· | ···················· |
| AB | 6393 | hCoV-19/England/CAMC-CB7831/2020\|EPI_ISL_777860\|20 | ···················· | ···················· | ···················· | ···················· | ···················· | ···················· |
| AB | 6394 | hCoV-19/England/CAMC-CB7DE7/2020\|EPI_ISL_777861\|20 | ···················· | ···················· | ···················· | ···················· | ···················· | ···················· |
| AB | 6395 | hCoV-19/England/CAMC-CB7A59/2020\|EPI_ISL_777862\|20 | ···················· | ···················· | ···················· | ···················· | ···················· | ···················· |

| | | | | | | | | |
|---|---|---|---|---|---|---|---|---|
| AB | 6396 | hCoV-19/England/CAMC-CB8979/2020\|EPI_ISL_777876\|20 | ···················· | ···················· | ···················· | ···················· | ···················· | ···················· |
| AB | 6397 | hCoV-19/England/CAMC-CB7B0B/2020\|EPI_ISL_777878\|20 | ···················· | ···················· | ···················· | ···················· | ···················· | ···················· |
| AB | 6398 | hCoV-19/England/CAMC-CB6B93/2020\|EPI_ISL_777881\|20 | ···················· | ···················· | ···················· | ···················· | ···················· | ···················· |
| AB | 6399 | hCoV-19/England/CAMC-CB8C61/2020\|EPI_ISL_777882\|20 | ···················· | ···················· | ···················· | ···················· | ···················· | ···················· |
| AB | 6400 | hCoV-19/England/CAMC-CB89A6/2020\|EPI_ISL_777893\|20 | ···················· | ···················· | ···················· | ···················· | ···················· | ···············R···· |
| AB | 6401 | hCoV-19/Netherlands/ZH-EMC-1125/2020\|EPI_ISL_76320 | ···················· | ···················· | ···················· | ···················· | ···················· | ···················· |
| AB | 6402 | hCoV-19/Netherlands/ZH-EMC-1126/2020\|EPI_ISL_76320 | ···················· | ···················· | ···················· | ···················· | ···················· | ···················· |
| AB | 6403 | hCoV-19/Netherlands/ZH-EMC-1127/2020\|EPI_ISL_76320 | ···················· | ···················· | ···················· | ···················· | ···················· | ···················· |
| AB | 6404 | hCoV-19/Netherlands/ZH-EMC-1133/2020\|EPI_ISL_76321 | ···················· | ···················· | ···················· | ···················· | ···················· | ···················· |
| AB | 6405 | hCoV-19/Netherlands/ZH-EMC-1137/2020\|EPI_ISL_76321 | ···················· | ···················· | ···················· | ···················· | ···················· | ···················· |
| AB | 6406 | hCoV-19/Netherlands/ZH-EMC-1188/2020\|EPI_ISL_76326 | ···················· | ···················· | ···················· | ···················· | ···················· | ···················· |
| AB | 6407 | hCoV-19/Netherlands/ZH-EMC-1189/2020\|EPI_ISL_76326 | ···················· | ···················· | ···················· | ···················· | ···················· | ···················· |
| AB | 6408 | hCoV-19/Netherlands/ZH-EMC-1190/2020\|EPI_ISL_76326 | ···················· | ···················· | ···················· | ···················· | ···················· | ···················· |
| AB | 6409 | hCoV-19/Netherlands/ZH-EMC-1193/2020\|EPI_ISL_76326 | ···················· | ···················· | ···················· | ···················· | ···················· | ···················· |
| AB | 6410 | hCoV-19/Netherlands/ZH-EMC-1194/2020\|EPI_ISL_76326 | ···················· | ···················· | ···················· | ···················· | ···················· | ···················· |
| AB | 6411 | hCoV-19/Netherlands/ZH-EMC-1195/2020\|EPI_ISL_76326 | ···················· | ···················· | ···················· | ···················· | ···················· | ···················· |
| AB | 6412 | hCoV-19/Netherlands/ZH-EMC-1196/2020\|EPI_ISL_76326 | ···················· | ···················· | ···················· | ···················· | ···················· | ···················· |
| AB | 6413 | hCoV-19/Netherlands/ZH-EMC-1197/2020\|EPI_ISL_76327 | ···················· | ···················· | ···················· | ···················· | ···················· | ···················· |
| AB | 6414 | hCoV-19/Netherlands/ZH-EMC-1199/2020\|EPI_ISL_76327 | ···················· | ···················· | ···················· | ···················· | ···················· | ···················· |
| AB | 6415 | hCoV-19/Netherlands/ZH-EMC-1201/2020\|EPI_ISL_76327 | ···················· | ···················· | ···················· | ···················· | ···················· | ···················· |
| AB | 6416 | hCoV-19/Netherlands/ZH-EMC-1202/2020\|EPI_ISL_76327 | ···················· | ···················· | ···················· | ···················· | ···················· | ···················· |
| AB | 6417 | hCoV-19/Netherlands/ZH-EMC-1203/2020\|EPI_ISL_76327 | ···················· | ···················· | ···················· | ···················· | ···················· | ···················· |
| AB | 6418 | hCoV-19/Netherlands/ZH-EMC-1204/2020\|EPI_ISL_76327 | ···················· | ···················· | ···················· | ···················· | ···················· | ···················· |
| AB | 6419 | hCoV-19/Netherlands/ZH-EMC-1205/2020\|EPI_ISL_76327 | ···················· | ···················· | ···················· | ···················· | ···················· | ···················· |
| AB | 6420 | hCoV-19/Netherlands/ZH-EMC-1206/2020\|EPI_ISL_76327 | ···················· | ···················· | ···················· | ···················· | ·····T·············· | ···················· |
| AB | 6421 | hCoV-19/Netherlands/ZH-EMC-1211/2020\|EPI_ISL_76328 | ···················· | ···················· | ···················· | ···················· | ···················· | ···················· |
| AB | 6422 | hCoV-19/Netherlands/ZH-EMC-1212/2020\|EPI_ISL_76328 | ···················· | ···················· | ···················· | ···················· | ···················· | ···················· |
| AB | 6423 | hCoV-19/Netherlands/ZH-EMC-1214/2020\|EPI_ISL_76328 | ···················· | ···················· | ···················· | ···················· | ···················· | ···················· |
| AB | 6424 | hCoV-19/Netherlands/ZH-EMC-1215/2020\|EPI_ISL_76328 | ···················· | ···················· | ···················· | ···················· | ···················· | ···················· |
| AB | 6425 | hCoV-19/Netherlands/ZH-EMC-1216/2020\|EPI_ISL_76328 | ···················· | ···················· | ···················· | ···················· | ···················· | ···················· |
| AB | 6426 | hCoV-19/Netherlands/ZH-EMC-1217/2020\|EPI_ISL_76328 | ···················· | ···················· | ···················· | ···················· | ···················· | ···················· |
| AB | 6427 | hCoV-19/Netherlands/ZH-EMC-1218/2020\|EPI_ISL_76328 | ···················· | ···················· | ···················· | ···················· | ···················· | ···················· |
| AB | 6428 | hCoV-19/Netherlands/ZH-EMC-1222/2020\|EPI_ISL_76329 | ···················· | ···················· | ···················· | ···················· | ···················· | ···················· |
| AB | 6429 | hCoV-19/Netherlands/ZH-EMC-1223/2020\|EPI_ISL_76329 | ···················· | ···················· | ···················· | ···················· | ···················· | ···················· |
| AB | 6430 | hCoV-19/Netherlands/ZH-EMC-1224/2020\|EPI_ISL_76329 | ···················· | ···················· | ···················· | ···················· | ···················· | ···················· |
| AB | 6431 | hCoV-19/Netherlands/ZH-EMC-1227/2020\|EPI_ISL_76329 | ···················· | ···················· | ···················· | ···················· | ···················· | ···················· |
| AB | 6432 | hCoV-19/Netherlands/ZH-EMC-1228/2020\|EPI_ISL_76329 | ···················· | ···················· | ···················· | ···················· | ···················· | ···················· |
| AB | 6433 | hCoV-19/Netherlands/ZH-EMC-1229/2020\|EPI_ISL_76329 | ···················· | ···················· | ···················· | ···················· | ···················· | ···················· |
| AB | 6434 | hCoV-19/Netherlands/ZH-EMC-1230/2020\|EPI_ISL_76329 | ···················· | ···················· | ···················· | ···················· | ···················· | ···················· |
| AB | 6435 | hCoV-19/England/OXON-F4076A/2020\|EPI_ISL_764535\|20 | ···················· | ···················· | ···················· | ···················· | ···················· | ···················· |
| AB | 6436 | hCoV-19/England/OXON-F40779/2020\|EPI_ISL_764536\|20 | ···················· | ···················· | ···················· | ···················· | ···················· | ···················· |
| AB | 6437 | hCoV-19/England/OXON-F4080D/2020\|EPI_ISL_764537\|20 | ···················· | ···················· | ···················· | ···················· | ···················· | ···················· |
| AB | 6438 | hCoV-19/Wales/OXON-F40858/2020\|EPI_ISL_764538\|2020 | ···················· | ···················· | ···················· | ···················· | ···················· | ···················· |
| AB | 6439 | hCoV-19/Northern Ireland/OXON-F40885/2020\|EPI_ISL_ | ···················· | ···················· | ···················· | ···················· | ···················· | ···················· |
| AB | 6440 | hCoV-19/England/OXON-F40919/2020\|EPI_ISL_764540\|20 | ···················· | ···················· | ···················· | ···················· | ···················· | ···················· |
| AB | 6441 | hCoV-19/Wales/OXON-F40982/2020\|EPI_ISL_764541\|2020 | ···················· | ···················· | ···················· | ···················· | ···················· | ···················· |
| AB | 6442 | hCoV-19/England/OXON-F409A0/2020\|EPI_ISL_764542\|20 | ···················· | ···················· | ···················· | ···················· | ···················· | ···················· |
| AB | 6443 | hCoV-19/England/OXON-F40A07/2020\|EPI_ISL_764543\|20 | ···················· | ···················· | ···················· | ···················· | ···················· | ···················· |
| AB | 6444 | hCoV-19/England/OXON-F40A9E/2020\|EPI_ISL_764544\|20 | ···················· | ···················· | ···················· | ···················· | ···················· | ···················· |
| AB | 6445 | hCoV-19/England/OXON-F40AAD/2020\|EPI_ISL_764545\|20 | ···················· | ···················· | ···················· | ···················· | ···················· | ···················· |
| AB | 6446 | hCoV-19/Wales/PHWC-4A394C/2020\|EPI_ISL_764590\|2020 | ···················· | ···················· | ···················· | ···················· | ···················· | ···················· |
| AB | 6447 | hCoV-19/Wales/PHWC-4A396A/2020\|EPI_ISL_764591\|2020 | ···················· | ···················· | ···················· | ···················· | ···················· | ···················· |
| AB | 6448 | hCoV-19/Wales/PHWC-4A3979/2020\|EPI_ISL_764592\|2020 | ···················· | ···················· | ···················· | ···················· | ···················· | ···················· |
| AB | 6449 | hCoV-19/Wales/PHWC-4A3997/2020\|EPI_ISL_764593\|2020 | ···················· | ···················· | ···················· | ···················· | ···················· | ···················· |
| AB | 6453 | hCoV-19/Wales/PHWC-4A3A49/2020\|EPI_ISL_764597\|2020 | ···················· | ···················· | ···················· | ···················· | ···················· | ···················· |
| AB | 6456 | hCoV-19/Wales/PHWC-4A3A85/2020\|EPI_ISL_764600\|2020 | ···················· | ···················· | ···················· | ···················· | ···················· | ···················· |
| AB | 6458 | hCoV-19/Wales/PHWC-4A3B73/2020\|EPI_ISL_764602\|2020 | ···················· | ···················· | ···················· | ···················· | ···················· | ···················· |
| AB | 6461 | hCoV-19/Wales/PHWC-4A3BCE/2020\|EPI_ISL_764605\|2020 | ···················· | ···················· | ···················· | ···················· | ···················· | ···················· |
| AB | 6462 | hCoV-19/Wales/PHWC-4A3BDD/2020\|EPI_ISL_764606\|2020 | ···················· | ···················· | ···················· | ···················· | ···················· | ···················· |
| AB | 6463 | hCoV-19/Wales/PHWC-4A3BEC/2020\|EPI_ISL_764607\|2020 | ···················· | ···················· | ···················· | ···················· | ···················· | ···················· |
| AB | 6464 | hCoV-19/Wales/PHWC-4A3C16/2020\|EPI_ISL_764608\|2020 | ···················· | ···················· | ···················· | ···················· | ···················· | ···················· |
| AB | 6465 | hCoV-19/Wales/PHWC-4A3C34/2020\|EPI_ISL_764609\|2020 | ···················· | ···················· | ···················· | ···················· | ···················· | ···················· |
| AB | 6466 | hCoV-19/Wales/PHWC-4A3C43/2020\|EPI_ISL_764610\|2020 | ···················· | ···················· | ···················· | ···················· | ···················· | ···················· |
| AB | 6467 | hCoV-19/Wales/PHWC-4A3C52/2020\|EPI_ISL_764611\|2020 | ···················· | ···················· | ···················· | ···················· | ···················· | ···················· |
| AB | 6468 | hCoV-19/Wales/PHWC-4A3C61/2020\|EPI_ISL_764612\|2020 | ···················· | ···················· | ···················· | ···················· | ···················· | ···················· |
| AB | 6469 | hCoV-19/Wales/PHWC-4A3C70/2020\|EPI_ISL_764613\|2020 | ···················· | ···················· | ···················· | ···················· | ···················· | ···················· |
| AB | 6470 | hCoV-19/Wales/PHWC-4A3C8F/2020\|EPI_ISL_764614\|2020 | ···················· | ···················· | ···················· | ···················· | ···················· | ···················· |
| AB | 6471 | hCoV-19/Wales/PHWC-4A3C9E/2020\|EPI_ISL_764615\|2020 | ···················· | ···················· | ···················· | ···················· | ···················· | ···················· |
| AB | 6472 | hCoV-19/Wales/PHWC-4A3CBC/2020\|EPI_ISL_764616\|2020 | ···················· | ···················· | ···················· | ···················· | ···················· | ···················· |
| AB | 6473 | hCoV-19/Wales/PHWC-4A3CCB/2020\|EPI_ISL_764617\|2020 | ···················· | ···················· | ···················· | ···················· | ···················· | ···················· |
| AB | 6474 | hCoV-19/Wales/PHWC-4A3CDA/2020\|EPI_ISL_764618\|2020 | ···················· | ···················· | ···················· | ···················· | ···················· | ···················· |
| AB | 6475 | hCoV-19/Wales/PHWC-4A3CE9/2020\|EPI_ISL_764619\|2020 | ···················· | ···················· | ···················· | ···················· | ···················· | ···················· |
| AB | 6476 | hCoV-19/Wales/PHWC-4A3CF8/2020\|EPI_ISL_764620\|2020 | ···················· | ···················· | ···················· | ···················· | ···················· | ···················· |
| AB | 6477 | hCoV-19/Wales/PHWC-4A3D04/2020\|EPI_ISL_764621\|2020 | ···················· | ···················· | ···················· | ···················· | ···················· | ···················· |
| AB | 6478 | hCoV-19/Wales/PHWC-4A3D13/2020\|EPI_ISL_764622\|2020 | ···················· | ···················· | ···················· | ···················· | ···················· | ···················· |
| AB | 6479 | hCoV-19/USA/MA-MGH-03507/2020\|EPI_ISL_765866\|2020- | ···················· | ···················· | ···················· | ···················· | ···················· | ···················· |
| AB | 6480 | hCoV-19/USA/MA-MGH-03512/2020\|EPI_ISL_765867\|2020- | ···················· | ···················· | ···················· | ···················· | ···················· | ···················· |
| AB | 6481 | hCoV-19/USA/MA-MGH-03517/2020\|EPI_ISL_765868\|2020- | ···················· | ···················· | ···················· | ···················· | ···················· | ···················· |
| AB | 6482 | hCoV-19/USA/MA-MGH-03519/2020\|EPI_ISL_765869\|2020- | ···················· | ···················· | ···················· | ···················· | ···················· | ···················· |
| AB | 6483 | hCoV-19/USA/MA-MGH-03522/2020\|EPI_ISL_765870\|2020- | ···················· | ···················· | ···················· | ···················· | ···················· | ···················· |

| | | | | | | | | |
|---|---|---|---|---|---|---|---|---|
| AB | 6484 | hCoV-19/USA/MA-MGH-03523/2020\|EPI_ISL_765871\|2020- | .................... | .................... | .................... | .................... | .................... | .................... |
| AB | 6485 | hCoV-19/USA/MA-MGH-03526/2020\|EPI_ISL_765872\|2020- | .................... | .................... | .................... | .................... | .................... | .................... |
| AB | 6486 | hCoV-19/USA/MA-MGH-03527/2020\|EPI_ISL_765873\|2020- | .................... | .................... | .................... | .................... | .................... | .................... |
| AB | 6487 | hCoV-19/USA/MA-MGH-03528/2020\|EPI_ISL_765874\|2020- | .................... | .................... | .................... | .................... | .................... | .................... |
| AB | 6488 | hCoV-19/USA/MA-MGH-03532/2020\|EPI_ISL_765876\|2020- | .................... | .................... | .................... | .................... | .................... | .................... |
| AB | 6489 | hCoV-19/USA/TX-DSHS-1943/2020\|EPI_ISL_765934\|2020- | .................... | .A.................. | .................... | .................... | .................... | .................... |
| AB | 6490 | hCoV-19/USA/TX-DSHS-1947/2020\|EPI_ISL_765935\|2020- | .................... | .A.................. | .................... | .................... | .................... | .................... |
| AB | 6491 | hCoV-19/USA/TX-DSHS-1952/2020\|EPI_ISL_765936\|2020- | .................... | .................... | .................... | .................... | .................... | .................... |
| AB | 6492 | hCoV-19/USA/TX-DSHS-1953/2020\|EPI_ISL_765937\|2020- | .................... | .A.................. | .................... | .................... | .................... | .................... |
| AB | 6493 | hCoV-19/USA/TX-DSHS-1954/2020\|EPI_ISL_765938\|2020- | .................... | .................... | .................... | .................... | .................... | .................... |
| AB | 6494 | hCoV-19/USA/TX-DSHS-1961/2020\|EPI_ISL_765939\|2020- | .................... | .................... | .................... | .................... | .................... | .................... |
| AB | 6495 | hCoV-19/USA/TX-DSHS-1967/2020\|EPI_ISL_765940\|2020- | .................... | .A.................. | .................... | .................... | .................... | .................... |
| AB | 6497 | hCoV-19/USA/AZ-WL-9900253/2020\|EPI_ISL_765948\|2020 | .................... | .................... | .................... | .................... | .................... | .................... |
| AB | 6498 | hCoV-19/USA/AZ-WL-CVG00039/2020\|EPI_ISL_765953\|202 | .................... | .A.................. | .................... | .................... | .................... | .................... |
| AB | 6499 | hCoV-19/USA/NY-Wadsworth-292662-01/2020\|EPI_ISL_76 | .................... | .................... | .................... | .................... | .................... | .................... |
| AB | 6500 | hCoV-19/USA/NY-Wadsworth-292664-01/2020\|EPI_ISL_76 | .................... | .................... | .................... | .................... | .................... | .................... |
| AB | 6501 | hCoV-19/USA/NY-Wadsworth-292665-01/2020\|EPI_ISL_76 | .................... | .................... | .................... | .................... | .................... | .................... |
| AB | 6502 | hCoV-19/USA/NY-Wadsworth-292678-01/2020\|EPI_ISL_76 | .................... | .A.................. | .................... | .................... | .................... | .................... |
| AB | 6503 | hCoV-19/USA/NY-Wadsworth-292679-01/2020\|EPI_ISL_76 | .................... | .A.................. | .................... | .................... | .................... | .................... |
| AB | 6504 | hCoV-19/USA/NY-Wadsworth-292680-01/2020\|EPI_ISL_76 | .................... | .A.................. | .................... | .................... | .................... | .................... |
| AB | 6505 | hCoV-19/USA/NY-Wadsworth-292681-01/2020\|EPI_ISL_76 | .................... | .................... | .................... | .................... | .................... | .................... |
| AB | 6506 | hCoV-19/USA/NY-Wadsworth-292682-01/2020\|EPI_ISL_76 | .................... | .A.................. | .................... | .................... | .................... | .................... |
| AB | 6507 | hCoV-19/USA/NY-Wadsworth-292685-01/2020\|EPI_ISL_76 | .................... | .................... | .................... | .................... | .................... | .................... |
| AB | 6508 | hCoV-19/USA/NY-Wadsworth-292686-01/2020\|EPI_ISL_76 | .................... | .................... | .................... | .................... | .................... | .................... |
| AB | 6509 | hCoV-19/USA/NY-Wadsworth-292702-01/2020\|EPI_ISL_76 | .................... | .................... | .................... | .................... | .................... | .................... |
| AB | 6510 | hCoV-19/USA/NY-Wadsworth-292704-01/2020\|EPI_ISL_76 | .................... | .................... | .................... | .................... | .................... | .................... |
| AB | 6511 | hCoV-19/USA/NY-Wadsworth-292705-01/2020\|EPI_ISL_76 | .................... | .................... | .................... | .................... | .................... | .................... |
| AB | 6512 | hCoV-19/USA/NY-Wadsworth-292731-01/2020\|EPI_ISL_76 | .................... | .................... | .................... | .................... | .................... | .................... |
| AB | 6513 | hCoV-19/USA/NY-Wadsworth-292733-01/2020\|EPI_ISL_76 | .................... | .................... | .................... | .................... | .................... | .................... |
| AB | 6514 | hCoV-19/USA/NY-Wadsworth-292734-01/2020\|EPI_ISL_76 | .................... | .................... | .................... | .................... | .................... | .................... |
| AB | 6515 | hCoV-19/Ireland/LK-NVRL-MPL24200/2020\|EPI_ISL_7677 | .................... | .................... | .................... | .................... | .................... | .................... |
| AB | 6516 | hCoV-19/Ireland/CN-NVRL-MPU68503/2020\|EPI_ISL_7677 | .................... | .................... | .................... | .................... | .................... | .................... |
| AB | 6518 | hCoV-19/Switzerland/SG-ETHZ-420663/2020\|EPI_ISL_76 | .................... | .................... | .................... | .................... | .................... | .................... |
| AB | 6522 | hCoV-19/Switzerland/SO-ETHZ-420732/2020\|EPI_ISL_76 | .................... | .................... | .................... | .................... | .................... | .................... |
| AB | 6524 | hCoV-19/Switzerland/SO-ETHZ-420725/2020\|EPI_ISL_76 | .................... | .................... | .................... | .................... | .................... | .................... |
| AB | 6525 | hCoV-19/Switzerland/SO-ETHZ-420736/2020\|EPI_ISL_76 | .................... | .................... | .................... | .................... | ..............KK.... | .................... |
| AB | 6526 | hCoV-19/Switzerland/SO-ETHZ-420487/2020\|EPI_ISL_76 | .................... | .................... | .................... | .................... | .................... | .................... |
| AB | 6528 | hCoV-19/England/QEUH-C5D85B/2020\|EPI_ISL_769402\|20 | .................... | .................... | .................... | .................... | .................... | .................... |
| AB | 6529 | hCoV-19/England/QEUH-C5C4D0/2020\|EPI_ISL_769405\|20 | .................... | .................... | .................... | .................... | .................... | .................... |
| AB | 6530 | hCoV-19/England/MILK-C609E9/2020\|EPI_ISL_769409\|20 | .................... | .................... | .................... | .................... | .................... | .................... |
| AB | 6531 | hCoV-19/England/QEUH-C5BA84/2020\|EPI_ISL_769413\|20 | .................... | .................... | .................... | .................... | .................... | ..A................. |
| AB | 6532 | hCoV-19/England/QEUH-C5BBDC/2020\|EPI_ISL_769418\|20 | .................... | .................... | .................... | .................... | .................... | .................... |
| AB | 6533 | hCoV-19/England/QEUH-C5BAEE/2020\|EPI_ISL_769419\|20 | .................... | .................... | .................... | .................... | .................... | .................... |
| AB | 6534 | hCoV-19/England/QEUH-C5D91C/2020\|EPI_ISL_769421\|20 | .................... | .................... | .................... | .................... | .................... | .................... |
| AB | 6535 | hCoV-19/England/QEUH-C5B987/2020\|EPI_ISL_769422\|20 | .................... | .................... | .................... | .................... | .................... | .................... |
| AB | 6536 | hCoV-19/England/MILK-C609BC/2020\|EPI_ISL_769423\|20 | .................... | .................... | .................... | .................... | .................... | .................... |
| AB | 6537 | hCoV-19/England/MILK-C633AC/2020\|EPI_ISL_769426\|20 | .................... | .................... | .................... | .................... | .................... | .................... |
| AB | 6538 | hCoV-19/England/QEUH-C5BE6A/2020\|EPI_ISL_769428\|20 | .................... | .................... | .................... | .................... | .................... | .................... |
| AB | 6539 | hCoV-19/England/MILK-C60C3B/2020\|EPI_ISL_769429\|20 | .................... | .................... | .................... | .................... | .................... | .................... |
| AB | 6540 | hCoV-19/England/MILK-C60C59/2020\|EPI_ISL_769432\|20 | .................... | .................... | .................... | .................... | .................... | .................... |
| AB | 6541 | hCoV-19/England/QEUH-C5C3F1/2020\|EPI_ISL_769434\|20 | .................... | .................... | .................... | .................... | .....T.............. | .................... |
| AB | 6542 | hCoV-19/England/MILK-C61915/2020\|EPI_ISL_769439\|20 | .................... | .................... | .................... | .................... | .................... | .................... |
| AB | 6543 | hCoV-19/England/MILK-C60CE0/2020\|EPI_ISL_769444\|20 | .................... | .................... | .................... | .................... | .................... | .................... |
| AB | 6544 | hCoV-19/England/QEUH-C5BDF4/2020\|EPI_ISL_769445\|20 | .................... | .................... | .................... | .................... | .................... | .................... |
| AB | 6545 | hCoV-19/England/MILK-C614BA/2020\|EPI_ISL_769447\|20 | .................... | .................... | .................... | .................... | .................... | ..A................. |
| AB | 6546 | hCoV-19/England/QEUH-C5BBFA/2020\|EPI_ISL_769450\|20 | .................... | .................... | .................... | .................... | .................... | .................... |
| AB | 6547 | hCoV-19/England/MILK-C60C68/2020\|EPI_ISL_769451\|20 | .................... | .................... | .................... | .................... | .................... | .................... |
| AB | 6548 | hCoV-19/England/QEUH-C5C027/2020\|EPI_ISL_769452\|20 | .................... | .................... | .................... | .................... | .................... | .................... |
| AB | 6549 | hCoV-19/England/MILK-C60BA7/2020\|EPI_ISL_769455\|20 | .................... | .................... | .................... | .................... | .................... | .................... |
| AB | 6550 | hCoV-19/England/QEUH-C5BEC4/2020\|EPI_ISL_769460\|20 | .................... | .................... | .................... | .................... | .................... | .................... |
| AB | 6551 | hCoV-19/England/MILK-C60C2C/2020\|EPI_ISL_769461\|20 | .................... | .................... | .................... | .................... | .................... | .................... |
| AB | 6552 | hCoV-19/England/MILK-C603BE/2020\|EPI_ISL_769463\|20 | .................... | .................... | .................... | .................... | .................... | .................... |
| AB | 6553 | hCoV-19/England/QEUH-C5BA0C/2020\|EPI_ISL_769470\|20 | .................... | .................... | .................... | .................... | .................... | .................... |
| AB | 6554 | hCoV-19/England/QEUH-C5BF94/2020\|EPI_ISL_769471\|20 | .................... | .................... | .................... | .................... | .................... | .................... |
| AB | 6555 | hCoV-19/England/QEUH-C5BFA3/2020\|EPI_ISL_769475\|20 | .................... | .................... | .................... | .................... | .................... | .................... |
| AB | 6556 | hCoV-19/England/MILK-C611C2/2020\|EPI_ISL_769477\|20 | .................... | .................... | .................... | .................... | .................... | .................... |
| AB | 6557 | hCoV-19/England/MILK-C613F9/2020\|EPI_ISL_769478\|20 | .................... | .................... | .................... | .................... | .................... | ..A................. |
| AB | 6558 | hCoV-19/England/MILK-C60D65/2020\|EPI_ISL_769480\|20 | .................... | .................... | .................... | .................... | .................... | .................... |
| AB | 6559 | hCoV-19/England/QEUH-C5BB81/2020\|EPI_ISL_769482\|20 | .................... | .................... | .................... | .................... | .................... | .................... |
| AB | 6560 | hCoV-19/England/MILK-C609DA/2020\|EPI_ISL_769483\|20 | .................... | .................... | .................... | .................... | .................... | .................... |
| AB | 6561 | hCoV-19/England/MILK-C60F32/2020\|EPI_ISL_769484\|20 | .................... | .................... | .................... | .................... | .................... | .................... |
| AB | 6562 | hCoV-19/England/MILK-C613BD/2020\|EPI_ISL_769490\|20 | .................... | .................... | .................... | .................... | .................... | .................... |
| AB | 6563 | hCoV-19/England/MILK-C6147E/2020\|EPI_ISL_769492\|20 | .................... | .................... | .................... | .................... | .................... | ..A................. |
| AB | 6564 | hCoV-19/England/QEUH-C5BFD0/2020\|EPI_ISL_769499\|20 | .................... | .................... | .................... | .................... | .................... | .................... |
| AB | 6565 | hCoV-19/USA/WY-WPHL-20171476/2020\|EPI_ISL_770444\|2 | .................... | .................... | .................... | .................... | .................... | .................... |
| AB | 6566 | hCoV-19/USA/WY-WPHL-20171626/2020\|EPI_ISL_770445\|2 | .................... | .................... | .................... | .................... | .................... | .................... |
| AB | 6567 | hCoV-19/USA/WY-WPHL-20171657/2020\|EPI_ISL_770447\|2 | .................... | .................... | .................... | .................... | .................... | .................... |
| AB | 6568 | hCoV-19/USA/WY-WPHL-20171663/2020\|EPI_ISL_770448\|2 | .................... | .A.................. | .................... | .................... | .................... | .................... |
| AB | 6569 | hCoV-19/USA/WY-WPHL-20171669/2020\|EPI_ISL_770449\|2 | .................... | .................... | .................... | .................... | .................... | .................... |
| AB | 6570 | hCoV-19/USA/WY-WPHL-20171712/2020\|EPI_ISL_770451\|2 | .................... | .................... | .................... | .................... | .................... | .................... |

| | | | | | | | |
|---|---|---|---|---|---|---|---|
| AB | 6571 hCoV-19/USA/WY-WPHL-20171797/2020\|EPI_ISL_770452\|2 | ···················· | ·A·················· | ···················· | ···················· | ···················· | ···················· |
| AB | 6572 hCoV-19/USA/WY-WPHL-20171805/2020\|EPI_ISL_770453\|2 | ···················· | ···················· | ···················· | ···················· | ···················· | ···················· |
| AB | 6573 hCoV-19/USA/WY-WPHL-20171884/2020\|EPI_ISL_770454\|2 | ···················· | ···················· | ···················· | ···················· | ···················· | ···················· |
| AB | 6574 hCoV-19/USA/WY-WPHL-20171946/2020\|EPI_ISL_770456\|2 | ···················· | ···················· | ···················· | ···················· | ···················· | ···················· |
| AB | 6575 hCoV-19/USA/WY-WPHL-20171966/2020\|EPI_ISL_770457\|2 | ···················· | ···················· | ···················· | ···················· | ···················· | ···················· |
| AB | 6576 hCoV-19/USA/WY-WPHL-20172216/2020\|EPI_ISL_770458\|2 | ···················· | ···················· | ···················· | ···················· | ···················· | ···················· |
| AB | 6577 hCoV-19/USA/WY-WPHL-20172249/2020\|EPI_ISL_770459\|2 | ···················· | ···················· | ···················· | ···················· | ···················· | ···················· |
| AB | 6578 hCoV-19/USA/WY-WPHL-20172332/2020\|EPI_ISL_770460\|2 | ···················· | ···················· | ···················· | ···················· | ···················· | ···················· |
| AB | 6579 hCoV-19/USA/WY-WPHL-20172560/2020\|EPI_ISL_770461\|2 | ···················· | ···················· | ···················· | ···················· | ···················· | ···················· |
| AB | 6581 hCoV-19/England/205300109/2020\|EPI_ISL_770467\|2020 | ···················· | ···················· | ···················· | ···················· | ···················· | N··················· |
| AB | 6583 hCoV-19/USA/WY-WPHL-20172245/2020\|EPI_ISL_770480\|2 | ···················· | ·A·················· | ···················· | ···················· | ···················· | ···················· |
| AB | 6584 hCoV-19/USA/WY-WPHL-20172338/2020\|EPI_ISL_770481\|2 | ···················· | ···················· | ···················· | ···················· | ···················· | ···················· |
| AB | 6585 hCoV-19/USA/WY-WPHL-20172364/2020\|EPI_ISL_770482\|2 | ···················· | ·A·················· | ···················· | ···················· | ···················· | ···················· |
| AB | 6586 hCoV-19/USA/WY-WPHL-20172399/2020\|EPI_ISL_770483\|2 | ···················· | ···················· | ···················· | ···················· | ···················· | ···················· |
| AB | 6587 hCoV-19/USA/WY-WPHL-20172411/2020\|EPI_ISL_770484\|2 | ···················· | ···················· | ···················· | ···················· | ···················· | ···················· |
| AB | 6588 hCoV-19/USA/WY-WPHL-20172449/2020\|EPI_ISL_770485\|2 | ···················· | ···················· | ···················· | ···················· | ···················· | ···················· |
| AB | 6589 hCoV-19/USA/WY-WPHL-20172467/2020\|EPI_ISL_770486\|2 | ···················· | ···················· | ···················· | ···················· | ···················· | ···················· |
| AB | 6590 hCoV-19/USA/WY-WPHL-20172536/2020\|EPI_ISL_770487\|2 | ···················· | ···················· | ···················· | ···················· | ···················· | ···················· |
| AB | 6591 hCoV-19/USA/WY-WPHL-20172574/2020\|EPI_ISL_770488\|2 | ···················· | ···················· | ···················· | ···················· | ···················· | ···················· |
| AB | 6592 hCoV-19/USA/WY-WPHL-20172604/2020\|EPI_ISL_770489\|2 | ···················· | ···················· | ···················· | ···················· | ···················· | ···················· |
| AB | 6593 hCoV-19/USA/WY-WPHL-20172609/2020\|EPI_ISL_770490\|2 | ···················· | ···················· | ···················· | ···················· | ···················· | ···················· |
| AB | 6594 hCoV-19/USA/WY-WPHL-20172644/2020\|EPI_ISL_770491\|2 | ···················· | ······A············· | ···················· | ···················· | ···················· | ···················· |
| AB | 6595 hCoV-19/USA/WY-WPHL-20172645/2020\|EPI_ISL_770492\|2 | ···················· | ···················· | ···················· | ···················· | ···················· | ···················· |
| AB | 6596 hCoV-19/USA/WY-WPHL-20172646/2020\|EPI_ISL_770493\|2 | ···················· | ···················· | ···················· | ···················· | ···················· | ···················· |
| AB | 6597 hCoV-19/USA/WY-WPHL-20172669/2020\|EPI_ISL_770494\|2 | ············T······· | ···················· | ···················· | ···················· | ···················· | ···················· |
| AB | 6598 hCoV-19/USA/WY-WPHL-20172741/2020\|EPI_ISL_770495\|2 | ···················· | ···················· | ···················· | ···················· | ···················· | ···················· |
| AB | 6599 hCoV-19/USA/WY-WPHL-21000114/2021\|EPI_ISL_770496\|2 | ···················· | ···················· | ···················· | ···················· | ···················· | ···················· |
| AB | 6600 hCoV-19/USA/WY-WPHL-21000146/2021\|EPI_ISL_770497\|2 | ···················· | ···················· | ···················· | ···················· | ···················· | ···················· |
| AB | 6601 hCoV-19/Lithuania/MR-LUHS-Eilnr237564/2020\|EPI_ISL | ···················· | ···················· | ···················· | ···················· | ···················· | ···················· |
| AB | 6602 hCoV-19/Lithuania/MR-LUHS-Eilnr572/2020\|EPI_ISL_77 | ···················· | ···················· | ···················· | ···················· | ···················· | ···················· |
| AB | 6603 hCoV-19/Morocco/FMP-51/2020\|EPI_ISL_775247\|2020-12 | ···················· | ···················· | ···················· | ···················· | ···················· | ···················· |
| AB | 6604 hCoV-19/Morocco/FMP-52/2020\|EPI_ISL_775249\|2020-12 | ···················· | ···················· | ···················· | ···················· | ···················· | ···················· |
| AB | 6605 hCoV-19/Morocco/FMP-53/2020\|EPI_ISL_775250\|2020-11 | ···················· | ···················· | ···················· | ···················· | ···················· | ···················· |
| AB | 6606 hCoV-19/Morocco/FMP-54/2020\|EPI_ISL_775252\|2020-11 | ···················· | ···················· | ···················· | ···················· | ···················· | ···················· |
| AB | 6607 hCoV-19/Morocco/FMP-55/2020\|EPI_ISL_775254\|2020-12 | ···················· | ···················· | ···················· | ···················· | ···················· | ···················· |
| AB | 6608 hCoV-19/Morocco/FMP-56/2020\|EPI_ISL_775255\|2020 | ···················· | ···················· | ···················· | ···················· | ···················· | ···················· |
| AB | 6609 hCoV-19/Morocco/FMP-57/2020\|EPI_ISL_775256\|2020-12 | ···················· | ···················· | ···················· | ···················· | ···················· | ···················· |
| AB | 6610 hCoV-19/Morocco/FMP-58/2020\|EPI_ISL_775258\|2020-12 | ···················· | ···················· | ···················· | ···················· | ···················· | ···················· |
| AB | 6611 hCoV-19/Italy/CAM-INTPas-20/2020\|EPI_ISL_775259\|20 | ···················· | ···················· | ···················· | ···················· | ···················· | ···················· |
| AB | 6612 hCoV-19/Morocco/FMP-59/2020\|EPI_ISL_775260\|2020-12 | ···················· | ···················· | ···················· | ···················· | ···················· | ···················· |
| AB | 6613 hCoV-19/Morocco/FMP-60/2020\|EPI_ISL_775262\|2020-12 | ···················· | ···················· | ···················· | ···················· | ···················· | ···················· |
| AB | 6614 hCoV-19/Morocco/FMP-61/2020\|EPI_ISL_775263\|2020-12 | ···················· | ·A·················· | ···················· | ···················· | ···················· | ··A················· |
| AB | 6615 hCoV-19/Morocco/FMP-62/2020\|EPI_ISL_775264\|2020-11 | ···················· | ···················· | ···················· | ···················· | ···················· | ···················· |
| AB | 6616 hCoV-19/Morocco/FMP-63/2020\|EPI_ISL_775265\|2020-11 | ···················· | ···················· | ···················· | ···················· | ···················· | ···················· |
| AB | 6617 hCoV-19/Norway/7021/2020\|EPI_ISL_775275\|2020-12-18 | ···················· | ···················· | ···················· | ···················· | ···················· | ···················· |
| AB | 6618 hCoV-19/Norway/6238/2020\|EPI_ISL_775276\|2020-11-25 | ···················· | ···················· | ···················· | ···················· | ···················· | ···················· |
| AB | 6619 hCoV-19/Norway/6595/2020\|EPI_ISL_775277\|2020-12-03 | ···················· | ·A·················· | ···················· | ···················· | ···················· | ···················· |
| AB | 6620 hCoV-19/Norway/7099/2020\|EPI_ISL_775278\|2020-12-17 | ···················· | ·A·················· | ···················· | ···················· | ···················· | ··A················· |
| AB | 6621 hCoV-19/Norway/4081/2020\|EPI_ISL_775279\|2020-10-03 | ···················· | ···················· | ···················· | ···················· | ···················· | ···················· |
| AB | 6622 hCoV-19/Norway/4561/2020\|EPI_ISL_775280\|2020-10-16 | ···················· | ···················· | ···················· | ···················· | ···················· | ···················· |
| AB | 6623 hCoV-19/Norway/5828/2020\|EPI_ISL_775281\|2020-11-05 | ···················· | ···················· | ···················· | ···················· | ···················· | ···················· |
| AB | 6624 hCoV-19/Norway/5829/2020\|EPI_ISL_775282\|2020-11-05 | ···················· | ···················· | ···················· | ···················· | ···················· | ···················· |
| AB | 6625 hCoV-19/Norway/5831/2020\|EPI_ISL_775283\|2020-11-05 | ···················· | ···················· | ···················· | ···················· | ···················· | ···················· |
| AB | 6626 hCoV-19/Norway/6132/2020\|EPI_ISL_775284\|2020-11-29 | ···················· | ···················· | ···················· | ···················· | ···················· | ···················· |
| AB | 6627 hCoV-19/Norway/6266/2020\|EPI_ISL_775285\|2020-11-30 | ···················· | ···················· | ···················· | ···················· | ···················· | ···················· |
| AB | 6628 hCoV-19/Norway/6273/2020\|EPI_ISL_775286\|2020-11-23 | ···················· | ···················· | ···················· | ···················· | ···················· | ···················· |
| AB | 6629 hCoV-19/Norway/6281/2020\|EPI_ISL_775287\|2020-11-24 | ···················· | ···················· | ···················· | ···················· | ···················· | ···················· |
| AB | 6630 hCoV-19/Norway/6286/2020\|EPI_ISL_775288\|2020-11-26 | ···················· | ···················· | ···················· | ···················· | ···················· | ···················· |
| AB | 6631 hCoV-19/Norway/6291/2020\|EPI_ISL_775289\|2020-11-25 | ···················· | ···················· | ···················· | ···················· | ···················· | ···················· |
| AB | 6632 hCoV-19/Norway/6294/2020\|EPI_ISL_775290\|2020-11-27 | ···················· | ···················· | ···················· | ···················· | ···················· | ···················· |
| AB | 6633 hCoV-19/Norway/6611/2020\|EPI_ISL_775291\|2020-12-03 | ···················· | ···················· | ···················· | ···················· | ···················· | ···················· |
| AB | 6634 hCoV-19/Norway/6614/2020\|EPI_ISL_775292\|2020-12-03 | ···················· | ···················· | ···················· | ···················· | ···················· | ···················· |
| AB | 6635 hCoV-19/Norway/6770/2020\|EPI_ISL_775293\|2020-12-09 | ···················· | ··G················· | ···················· | ···················· | ···················· | ···················· |
| AB | 6636 hCoV-19/Norway/5823/2020\|EPI_ISL_775294\|2020-11-19 | ···················· | ···················· | ···················· | ···················· | ···················· | ···················· |
| AB | 6637 hCoV-19/Norway/5832/2020\|EPI_ISL_775295\|2020-11-03 | ···················· | ···················· | ···················· | ···················· | ···················· | ···················· |
| AB | 6638 hCoV-19/Norway/5833/2020\|EPI_ISL_775296\|2020-11-05 | ···················· | ···················· | ···················· | ···················· | ···················· | ···················· |
| AB | 6639 hCoV-19/Norway/5834/2020\|EPI_ISL_775297\|2020-11-17 | ···················· | ···················· | ···················· | ···················· | ···················· | ···················· |
| AB | 6640 hCoV-19/Norway/5835/2020\|EPI_ISL_775298\|2020-11-13 | ···················· | ···················· | ···················· | ···················· | ···················· | ···················· |
| AB | 6641 hCoV-19/Norway/5839/2020\|EPI_ISL_775299\|2020-11-06 | ···················· | ···················· | ···················· | ···················· | ···················· | ···················· |
| AB | 6642 hCoV-19/Germany/HH-hpi-p2447/2020\|EPI_ISL_776510\|2 | ···················· | ···················· | ···················· | ···················· | ···················· | ···················· |
| AB | 6644 hCoV-19/Germany/HH-hpi-p2450/2020\|EPI_ISL_776512\|2 | ···················· | ···················· | ···················· | ···················· | ···················· | ···················· |
| AB | 6645 hCoV-19/Germany/HH-hpi-p2451/2020\|EPI_ISL_776513\|2 | ···················· | ···················· | ···················· | ···················· | ···················· | ···················· |
| AB | 6646 hCoV-19/England/CAMC-CB7C17/2020\|EPI_ISL_777841\|20 | ···················· | ···················· | ···················· | ···················· | ···················· | ···················· |
| AB | 6647 hCoV-19/Spain/M-20115992/2020\|EPI_ISL_777902\|2020- | ···················· | ···················· | ···················· | ···················· | ···················· | ···················· |
| AB | 6648 hCoV-19/England/CAMC-CB88F4/2020\|EPI_ISL_777904\|20 | ···················· | ···················· | ···················· | ···················· | ···················· | ···················· |
| AB | 6649 hCoV-19/England/CAMC-CB753A/2020\|EPI_ISL_777906\|20 | ···················· | ···················· | ···················· | ···················· | ······T············· | ···················· |
| AB | 6650 hCoV-19/England/CAMC-CB8C34/2020\|EPI_ISL_777910\|20 | ···················· | ···················· | ···················· | ···················· | ···················· | ···················· |
| AB | 6651 hCoV-19/England/CAMC-CB7EC6/2020\|EPI_ISL_777914\|20 | ···················· | ···················· | ···················· | ···················· | ···················· | ···················· |
| AB | 6652 hCoV-19/England/CAMC-CB8672/2020\|EPI_ISL_777916\|20 | ···················· | ···················· | ···················· | ···················· | ···················· | ···················· |
| AB | 6653 hCoV-19/England/CAMC-CB8BCE/2020\|EPI_ISL_777919\|20 | ···················· | ···················· | ···················· | ···················· | ···················· | ···················· |

```
AB    6654 hCoV-19/England/CAMC-CB8706/2020|EPI_ISL_777923|20    ····················  ····················  ·····················  ····················  ····················  ····················
AB    6655 hCoV-19/England/CAMC-CB79C5/2020|EPI_ISL_777924|20    ····················  ····················  ·····················  ····················  ····················  ····················
AB    6656 hCoV-19/England/CAMC-CB8B28/2020|EPI_ISL_777925|20    ···········T········  ····················  ·····················  ····················  ····················  ····················
AB    6657 hCoV-19/England/ALDP-CBF4F9/2020|EPI_ISL_760267|20    ····················  ····················  ·····················  ····················  ····················  ····················
AB    6658 hCoV-19/England/ALDP-CC0914/2020|EPI_ISL_760268|20    ····················  ····················  ·····················  ····················  ····················  ···A················
AB    6659 hCoV-19/England/QEUH-CC594C/2020|EPI_ISL_760269|20    ····················  ····················  ·····················  ····················  ····················  ····················
AB    6660 hCoV-19/England/QEUH-CC2657/2020|EPI_ISL_760272|20    ····················  ····················  ·····················  ····················  ····················  ····················
AB    6661 hCoV-19/England/QEUH-CC5DC8/2020|EPI_ISL_760275|20    ····················  ····················  ·····················  ····················  ····················  ····················
AB    6662 hCoV-19/England/QEUH-CC300D/2020|EPI_ISL_760277|20    ····················  ····················  ·····················  ····················  ····················  ····················
AB    6663 hCoV-19/England/QEUH-CC2332/2020|EPI_ISL_760279|20    ····················  ····················  ·····················  ····················  ····················  ····················
AB    6664 hCoV-19/England/QEUH-CC778C/2020|EPI_ISL_760280|20    ····················  ····················  ·····················  ····················  ····················  ····················
AB    6665 hCoV-19/England/QEUH-CC2FC5/2020|EPI_ISL_760283|20    ····················  ····················  ·····················  ····················  ····················  ····················
AB    6666 hCoV-19/Wales/ALDP-CC089F/2020|EPI_ISL_760284|2020    ····················  ····················  ·····················  ····················  ····················  ····················
AB    6667 hCoV-19/England/QEUH-CC2675/2020|EPI_ISL_760289|20    ····················  ····················  ·····················  ····················  ····················  ····················
AB    6668 hCoV-19/England/ALDP-CBE023/2020|EPI_ISL_760293|20    ····················  ····················  ·····················  ····················  ····················  ····················
AB    6669 hCoV-19/England/ALDP-CBE041/2020|EPI_ISL_760296|20    ····················  ····················  ·····················  ····················  ····················  ····················
AB    6670 hCoV-19/England/MILK-CA63CB/2020|EPI_ISL_761599|20    ····················  ····················  ·····················  ····················  ····················  ····················
AB    6672 hCoV-19/Wales/PHWC-4A6AEC/2020|EPI_ISL_765101|2020    ····················  ····················  ·····················  ····················  ····················  ····················
AB    6673 hCoV-19/Wales/PHWC-4A6B07/2020|EPI_ISL_765102|2020    ····················  ····················  ·····················  ····················  ····················  ····················
AB    6675 hCoV-19/Wales/PHWC-4A6B43/2020|EPI_ISL_765104|2020    ····················  ····················  ·····················  ····················  ····················  ····················
AB    6676 hCoV-19/Wales/PHWC-4A6B70/2020|EPI_ISL_765105|2020    ····················  ····················  ·····················  ····················  ····················  ····················
AB    6678 hCoV-19/Wales/PHWC-4A6C22/2020|EPI_ISL_765110|2020    ····················  ····················  ·····················  ····················  ····················  ····················
AB    6679 hCoV-19/England/PORT-2D215D/2020|EPI_ISL_765112|20    ····················  ····················  ·····················  ····················  ····················  ····················
AB    6680 hCoV-19/England/PORT-2D258E/2020|EPI_ISL_765113|20    ····················  ····················  ·····················  ····················  ····················  ····················
AB    6681 hCoV-19/England/PORT-2D25F7/2020|EPI_ISL_765114|20    ····················  ····················  ·····················  ····················  ····················  ····················
AB    6682 hCoV-19/England/PORT-2D5460/2020|EPI_ISL_765115|20    ····················  ····················  ·····················  ····················  ····················  ····················
AB    6683 hCoV-19/England/PORT-2D549D/2020|EPI_ISL_765116|20    ····················  ·········T··········  ·····················  ····················  ····················  ····················
AB    6684 hCoV-19/England/PORT-2D54AC/2020|EPI_ISL_765117|20    ····················  ····················  ·····················  ····················  ······T·············  ····················
AB    6685 hCoV-19/England/PORT-2D54E8/2020|EPI_ISL_765118|20    ····················  ····················  ·····················  ····················  ······T·············  ····················
AB    6686 hCoV-19/England/PORT-2D54F7/2020|EPI_ISL_765119|20    ····················  ····················  ·····················  ····················  ····················  ····················
AB    6687 hCoV-19/England/PORT-2D5521/2020|EPI_ISL_765120|20    ····················  ····················  ·····················  ····················  ····················  ····················
AB    6688 hCoV-19/England/PORT-2D5530/2020|EPI_ISL_765121|20    ····················  ····················  ·····················  ····················  ····················  ····················
AB    6689 hCoV-19/England/PORT-2D559A/2020|EPI_ISL_765122|20    ····················  ····················  ·····················  ····················  ····················  ····················
AB    6690 hCoV-19/England/PORT-2D566A/2020|EPI_ISL_765123|20    ····················  ····················  ·····················  ····················  ····················  ····················
AB    6692 hCoV-19/England/PORT-2D5697/2020|EPI_ISL_765125|20    ····················  ····················  ·····················  ····················  ····················  ····················
AB    6693 hCoV-19/England/PORT-2D56C4/2020|EPI_ISL_765126|20    ····················  ····················  ·····················  ····················  ····················  ····················
AB    6694 hCoV-19/England/PORT-2D571C/2020|EPI_ISL_765127|20    ····················  ····················  ·····················  ····················  ······T·············  ····················
AB    6695 hCoV-19/England/PORT-2D5794/2020|EPI_ISL_765128|20    ····················  ····················  ·····················  ····················  ····················  ····················
AB    6696 hCoV-19/England/PORT-2D57C1/2020|EPI_ISL_765129|20    ····················  ····················  ·····················  ····················  ····················  ····················
AB    6697 hCoV-19/England/PORT-2D57D0/2020|EPI_ISL_765130|20    ····················  ····················  ·····················  ····················  ····················  ····················
AB    6698 hCoV-19/England/PORT-2D5916/2020|EPI_ISL_765131|20    ····················  ····················  ·····················  ····················  ····················  ····················
AB    6699 hCoV-19/England/PORT-2D5925/2020|EPI_ISL_765132|20    ····················  ····················  ·····················  ····················  ····················  ····················
AB    6700 hCoV-19/England/PORT-2D598F/2020|EPI_ISL_765133|20    ····················  ····················  ·····················  ····················  ····················  ····················
AB    6701 hCoV-19/England/PORT-2D59CB/2020|EPI_ISL_765134|20    ····················  ····················  ·····················  ····················  ····················  ····················
AB    6702 hCoV-19/England/PORT-2D5AE6/2020|EPI_ISL_765135|20    ····················  ····················  ·····················  ····················  ····················  ····················
AB    6703 hCoV-19/England/PORT-2D5AF5/2020|EPI_ISL_765136|20    ····················  ····················  ·····················  ····················  ······T·············  ····················
AB    6704 hCoV-19/England/PORT-2D5B7A/2020|EPI_ISL_765137|20    ····················  ····················  ·····················  ····················  ····················  ····················
AB    6705 hCoV-19/England/PORT-2D5BD4/2020|EPI_ISL_765138|20    ····················  ····················  ·····················  ····················  ····················  ····················
AB    6706 hCoV-19/England/PORT-2D5C3B/2020|EPI_ISL_765139|20    ····················  ····················  ·····················  ····················  ····················  ····················
AB    6707 hCoV-19/England/PORT-2D5C86/2020|EPI_ISL_765140|20    ····················  ····················  ·····················  ····················  ····················  ····················
AB    6708 hCoV-19/England/PORT-2D5CA4/2020|EPI_ISL_765141|20    ····················  ····················  ·····················  ····················  ······T·············  ····················
AB    6709 hCoV-19/England/PORT-2D5D0B/2020|EPI_ISL_765142|20    ····················  ····················  ·····················  ····················  ····················  ····················
AB    6710 hCoV-19/England/PORT-2D5D74/2020|EPI_ISL_765143|20    ····················  ····················  ·····················  ····················  ····················  ····················
AB    6711 hCoV-19/England/PORT-2D5D83/2020|EPI_ISL_765144|20    ····················  ····················  ·····················  ····················  ····················  ····················
AB    6712 hCoV-19/England/PORT-2D5DFC/2020|EPI_ISL_765145|20    ····················  ····················  ·····················  ····················  ····················  ····················
AB    6713 hCoV-19/England/PORT-2D5E08/2020|EPI_ISL_765146|20    ····················  ····················  ·····················  ····················  ······T·············  ····················
AB    6714 hCoV-19/England/PORT-2EE710/2020|EPI_ISL_765147|20    ····················  ····················  ·····················  ····················  ····················  ····················
AB    6715 hCoV-19/England/PORT-2EE7C5/2020|EPI_ISL_765148|20    ····················  ····················  ·····················  ····················  ····················  ····················
AB    6716 hCoV-19/England/SHEF-10C5963/2020|EPI_ISL_765149|2    ····················  ····················  ·····················  ····················  ····················  ····················
AB    6717 hCoV-19/England/SHEF-10C5E19/2020|EPI_ISL_765150|2    ····················  ····················  ·····················  ····················  ····················  ····················
AB    6718 hCoV-19/England/SHEF-10C611F/2020|EPI_ISL_765151|2    ····················  ····················  ·····················  ····················  ····················  ····················
AB    6719 hCoV-19/England/SHEF-10C65B9/2020|EPI_ISL_765152|2    ····················  ····················  ·····················  ····················  ····················  ····················
AB    6720 hCoV-19/England/SHEF-10C6698/2020|EPI_ISL_765153|2    ····················  ····················  ·····················  ····················  ····················  ····················
AB    6721 hCoV-19/England/SHEF-10C6DEE/2020|EPI_ISL_765154|2    ····················  ····················  ·····················  ····················  ····················  ····················
AB    6722 hCoV-19/England/SHEF-10C762E/2020|EPI_ISL_765155|2    ····················  ····················  ·····················  ····················  ····················  ····················
AB    6723 hCoV-19/England/SHEF-10C7819/2020|EPI_ISL_765156|2    ····················  ····················  ·····················  ····················  ····················  ····················
AB    6724 hCoV-19/England/SHEF-10C7CA4/2020|EPI_ISL_765157|2    ····················  ····················  ·····················  ····················  ····················  ····················
AB    6725 hCoV-19/England/SHEF-10C8D55/2020|EPI_ISL_765158|2    ····················  ····················  ·····················  ····················  ····················  ····················
AB    6726 hCoV-19/England/SHEF-10C8F9B/2020|EPI_ISL_765159|2    ····················  ·N··················  ·····················  ····················  ····················  ···A················
AB    6727 hCoV-19/England/205140701/2020|EPI_ISL_766400|2020    ····················  ····················  ·····················  ····················  ····················  ····················
AB    6728 hCoV-19/England/205140703/2020|EPI_ISL_766401|2020    ····················  ····················  ·····················  ····················  ····················  ····················
AB    6729 hCoV-19/England/205140900/2020|EPI_ISL_766402|2020    ····················  ····················  ·····················  ····················  ····················  ····················
AB    6730 hCoV-19/England/205140903/2020|EPI_ISL_766403|2020    ····················  ····················  ·····················  ····················  ····················  ····················
AB    6731 hCoV-19/England/205140908/2020|EPI_ISL_766404|2020    ····················  ····················  ·····················  ····················  ····················  ····················
AB    6732 hCoV-19/England/205140909/2020|EPI_ISL_766405|2020    ····················  ····················  ·····················  ····················  ····················  ····················
AB    6733 hCoV-19/England/205140910/2020|EPI_ISL_766406|2020    ····················  ····················  ·····················  ····················  ····················  ····················
AB    6734 hCoV-19/England/205140913/2020|EPI_ISL_766407|2020    ····················  ····················  ·····················  ····················  ····················  ····················
AB    6735 hCoV-19/England/205140963/2020|EPI_ISL_766408|2020    ····················  ····················  ·····················  ····················  ····················  ····················
AB    6736 hCoV-19/England/205140964/2020|EPI_ISL_766409|2020    ····················  ····················  ·····················  ····················  ······T·············  ····················
AB    6737 hCoV-19/England/205141528/2020|EPI_ISL_766410|2020    ····················  ····················  ·····················  ····················  ····················  ····················
```

| | | | | | | | | |
|---|---|---|---|---|---|---|---|---|
| AB | 6738 | hCoV-19/England/205141537/2020\|EPI_ISL_766411\|2020 | ···················· | ···················· | ···················· | ···················· | ···················· | ···················· |
| AB | 6739 | hCoV-19/England/205141710/2020\|EPI_ISL_766412\|2020 | ···················· | ···················· | ···················· | ···················· | ···················· | ···················· |
| AB | 6740 | hCoV-19/England/205141733/2020\|EPI_ISL_766413\|2020 | ···················· | ···················· | ···················· | ···················· | ···················· | ···················· |
| AB | 6741 | hCoV-19/England/205141760/2020\|EPI_ISL_766414\|2020 | ···················· | ···················· | ···················· | ···················· | ···················· | ···················· |
| AB | 6742 | hCoV-19/England/205141859/2020\|EPI_ISL_766415\|2020 | ···················· | ···················· | ···················· | ···················· | ···················· | ···················· |
| AB | 6743 | hCoV-19/England/205160431/2020\|EPI_ISL_766416\|2020 | ···················· | ···················· | ···················· | ···················· | ···················· | ···················· |
| AB | 6744 | hCoV-19/England/205160433/2020\|EPI_ISL_766417\|2020 | ···················· | ···················· | ···················· | ···················· | ···················· | ···················· |
| AB | 6745 | hCoV-19/England/205160437/2020\|EPI_ISL_766418\|2020 | ···················· | ···················· | ···················· | ···················· | ···················· | ···················· |
| AB | 6746 | hCoV-19/England/205160470/2020\|EPI_ISL_766419\|2020 | ···················· | ···················· | ···················· | ···················· | ···················· | ···················· |
| AB | 6747 | hCoV-19/England/205160487/2020\|EPI_ISL_766420\|2020 | ···················· | ···················· | ···················· | ···················· | ···················· | ···················· |
| AB | 6748 | hCoV-19/England/205160488/2020\|EPI_ISL_766421\|2020 | ···················· | ···················· | ···················· | ···················· | ···················· | ···················· |
| AB | 6749 | hCoV-19/England/205160491/2020\|EPI_ISL_766422\|2020 | ···················· | ···················· | ···················· | ···················· | ···················· | ···················· |
| AB | 6750 | hCoV-19/England/205160492/2020\|EPI_ISL_766423\|2020 | ···················· | ···················· | ···················· | ···················· | ···················· | ···················· |
| AB | 6751 | hCoV-19/England/205160918/2020\|EPI_ISL_766424\|2020 | ···················· | ···················· | ···················· | ···················· | ···················· | ···················· |
| AB | 6752 | hCoV-19/England/205160929/2020\|EPI_ISL_766425\|2020 | ···················· | ···················· | ···················· | ···················· | ···················· | ···················· |
| AB | 6753 | hCoV-19/England/205160985/2020\|EPI_ISL_766426\|2020 | ···················· | ···················· | ···················· | ···················· | ···················· | ···················· |
| AB | 6754 | hCoV-19/England/205160989/2020\|EPI_ISL_766427\|2020 | ···················· | ···················· | ···················· | ···················· | ···················· | ···················· |
| AB | 6755 | hCoV-19/England/205160992/2020\|EPI_ISL_766428\|2020 | ···················· | ···················· | ···················· | ···················· | ···················· | ···················· |
| AB | 6756 | hCoV-19/England/205160999/2020\|EPI_ISL_766429\|2020 | ···················· | ···················· | ···················· | ···················· | ···················· | ···A················ |
| AB | 6757 | hCoV-19/England/205161120/2020\|EPI_ISL_766430\|2020 | ···················· | ···················· | ···················· | ···················· | ···················· | ···················· |
| AB | 6758 | hCoV-19/England/205161237/2020\|EPI_ISL_766431\|2020 | ···················· | ···················· | ···················· | ···················· | ···················· | ···················· |
| AB | 6759 | hCoV-19/England/205161239/2020\|EPI_ISL_766432\|2020 | ···················· | ···················· | ···················· | ···················· | ···················· | ···················· |
| AB | 6760 | hCoV-19/England/205161373/2020\|EPI_ISL_766433\|2020 | ···················· | ···················· | ···················· | ···················· | ···················· | ···················· |
| AB | 6761 | hCoV-19/England/205161440/2020\|EPI_ISL_766434\|2020 | ···················· | ···················· | ···················· | ···················· | ···················· | ···················· |
| AB | 6762 | hCoV-19/England/205161451/2020\|EPI_ISL_766435\|2020 | ···················· | ···················· | ···················· | ···················· | ···················· | ···················· |
| AB | 6763 | hCoV-19/England/205161452/2020\|EPI_ISL_766436\|2020 | ···················· | ···················· | ···················· | ···················· | ···················· | ···················· |
| AB | 6764 | hCoV-19/England/205161454/2020\|EPI_ISL_766437\|2020 | ···················· | ···················· | ···················· | ···················· | ···················· | ···················· |
| AB | 6765 | hCoV-19/England/205161562/2020\|EPI_ISL_766440\|2020 | ···················· | ···················· | ···················· | ···················· | ···················· | ···················· |
| AB | 6766 | hCoV-19/England/205161623/2020\|EPI_ISL_766442\|2020 | ···················· | ···················· | ···················· | ···················· | ···················· | ···················· |
| AB | 6767 | hCoV-19/England/205161624/2020\|EPI_ISL_766443\|2020 | ···················· | ···················· | ···················· | ···················· | ···················· | ···················· |
| AB | 6768 | hCoV-19/England/205161626/2020\|EPI_ISL_766444\|2020 | ···················· | ···················· | ···················· | ···················· | ···················· | ···················· |
| AB | 6769 | hCoV-19/England/205161631/2020\|EPI_ISL_766445\|2020 | ···················· | ···················· | ···················· | ···················· | ···················· | ···················· |
| AB | 6770 | hCoV-19/England/205161633/2020\|EPI_ISL_766446\|2020 | ···················· | ···················· | ···················· | ···················· | ···················· | ···················· |
| AB | 6771 | hCoV-19/England/205161648/2020\|EPI_ISL_766447\|2020 | ···················· | ···················· | ···················· | ···················· | ···················· | ···················· |
| AB | 6772 | hCoV-19/England/205180033/2020\|EPI_ISL_766448\|2020 | ···················· | ···················· | ···················· | ···················· | ···················· | ···················· |
| AB | 6773 | hCoV-19/England/205180276/2020\|EPI_ISL_766449\|2020 | ···················· | ···················· | ···················· | ···················· | ···················· | ···················· |
| AB | 6774 | hCoV-19/England/205180357/2020\|EPI_ISL_766450\|2020 | ···················· | ···················· | ···················· | ···················· | ···················· | ···················· |
| AB | 6775 | hCoV-19/England/205180359/2020\|EPI_ISL_766451\|2020 | ···················· | ···················· | ···················· | ···················· | ···················· | ···················· |
| AB | 6776 | hCoV-19/England/205180367/2020\|EPI_ISL_766452\|2020 | ···················· | ···················· | ···················· | ···················· | ···················· | ···················· |
| AB | 6777 | hCoV-19/England/205180413/2020\|EPI_ISL_766453\|2020 | ···················· | ···················· | ···················· | ···················· | ···················· | ···················· |
| AB | 6778 | hCoV-19/England/205180592/2020\|EPI_ISL_766454\|2020 | ···················· | ···················· | ···················· | ···················· | ···················· | ···················· |
| AB | 6779 | hCoV-19/England/205180593/2020\|EPI_ISL_766455\|2020 | ···················· | ···················· | ···················· | ···················· | ···················· | ···················· |
| AB | 6780 | hCoV-19/England/205180597/2020\|EPI_ISL_766456\|2020 | ···················· | ···················· | ···················· | ···················· | ···················· | ···················· |
| AB | 6781 | hCoV-19/England/205180603/2020\|EPI_ISL_766457\|2020 | ···················· | ···················· | ···················· | ···················· | ···················· | ···················· |
| AB | 6782 | hCoV-19/England/205180724/2020\|EPI_ISL_766458\|2020 | ···················· | ···················· | ···················· | ···················· | ···················· | ···················· |
| AB | 6783 | hCoV-19/England/205180725/2020\|EPI_ISL_766459\|2020 | ···················· | ···················· | ···················· | ···················· | ···················· | ···················· |
| AB | 6784 | hCoV-19/England/205180733/2020\|EPI_ISL_766460\|2020 | ···················· | ···················· | ···················· | ···················· | ···················· | ···················· |
| AB | 6785 | hCoV-19/England/205180734/2020\|EPI_ISL_766461\|2020 | ···················· | ···················· | ···················· | ···················· | ···················· | ···················· |
| AB | 6786 | hCoV-19/England/205180736/2020\|EPI_ISL_766462\|2020 | ···················· | ···················· | ···················· | ···················· | ···················· | ···················· |
| AB | 6787 | hCoV-19/England/205180743/2020\|EPI_ISL_766463\|2020 | ···················· | ···················· | ···················· | ···················· | ···················· | ···················· |
| AB | 6788 | hCoV-19/England/205180745/2020\|EPI_ISL_766464\|2020 | ···················· | ···················· | ···················· | ···················· | ···················· | ···················· |
| AB | 6789 | hCoV-19/England/205180777/2020\|EPI_ISL_766465\|2020 | ···················· | ···················· | ···················· | ···················· | ···················· | ···················· |
| AB | 6790 | hCoV-19/England/205180782/2020\|EPI_ISL_766466\|2020 | ···················· | ···················· | ···················· | ···················· | ···················· | ···················· |
| AB | 6791 | hCoV-19/England/205180788/2020\|EPI_ISL_766467\|2020 | ···················· | ···················· | ···················· | ···················· | ···················· | ···················· |
| AB | 6792 | hCoV-19/England/205180792/2020\|EPI_ISL_766468\|2020 | ···················· | ···················· | ···················· | ···················· | ···················· | ···················· |
| AB | 6793 | hCoV-19/England/205180794/2020\|EPI_ISL_766469\|2020 | ···················· | ···················· | ···················· | ···················· | ···················· | ···················· |
| AB | 6794 | hCoV-19/England/205180796/2020\|EPI_ISL_766470\|2020 | ···················· | ···················· | ···················· | ···················· | ···················· | ···················· |
| AB | 6795 | hCoV-19/England/205180824/2020\|EPI_ISL_766471\|2020 | ···················· | ···················· | ···················· | ···················· | ···················· | ···················· |
| AB | 6796 | hCoV-19/England/205180826/2020\|EPI_ISL_766472\|2020 | ···················· | ···················· | ···················· | ···················· | ···················· | ···················· |
| AB | 6797 | hCoV-19/England/205180834/2020\|EPI_ISL_766473\|2020 | ···················· | ···················· | ···················· | ···················· | ···················· | ···················· |
| AB | 6798 | hCoV-19/England/205181315/2020\|EPI_ISL_766474\|2020 | ···················· | ···················· | ···················· | ···················· | ···················· | ···················· |
| AB | 6799 | hCoV-19/England/205181397/2020\|EPI_ISL_766475\|2020 | ···················· | ···················· | ···················· | ···················· | ···················· | ···················· |
| AB | 6800 | hCoV-19/England/205181406/2020\|EPI_ISL_766476\|2020 | ···················· | ···················· | ···················· | ···················· | ···················· | ···················· |
| AB | 6801 | hCoV-19/England/205181570/2020\|EPI_ISL_766477\|2020 | ···················· | ···················· | ···················· | ···················· | ···················· | ···················· |
| AB | 6802 | hCoV-19/England/205181819/2020\|EPI_ISL_766478\|2020 | ···················· | ···················· | ···················· | ···················· | ···················· | ···················· |
| AB | 6803 | hCoV-19/England/205181828/2020\|EPI_ISL_766479\|2020 | ···················· | ···················· | ···················· | ···················· | ···················· | ···················· |
| AB | 6804 | hCoV-19/England/205181831/2020\|EPI_ISL_766480\|2020 | ···················· | ···················· | ···················· | ···················· | ···················· | ···················· |
| AB | 6805 | hCoV-19/England/205181927/2020\|EPI_ISL_766481\|2020 | ···················· | ···················· | ···················· | ···················· | ···················· | ···················· |
| AB | 6806 | hCoV-19/England/205181928/2020\|EPI_ISL_766482\|2020 | ···················· | ···················· | ···················· | ···················· | ···················· | ···················· |
| AB | 6807 | hCoV-19/England/205181929/2020\|EPI_ISL_766483\|2020 | ···················· | ···················· | ···················· | ···················· | ···················· | ···················· |
| AB | 6808 | hCoV-19/England/205181930/2020\|EPI_ISL_766484\|2020 | ···················· | ···················· | ···················· | ···················· | ···················· | ···················· |
| AB | 6809 | hCoV-19/England/205181931/2020\|EPI_ISL_766485\|2020 | ···················· | ···················· | ···················· | ···················· | ···················· | ···················· |
| AB | 6810 | hCoV-19/England/205181932/2020\|EPI_ISL_766486\|2020 | ···················· | ···················· | ···················· | ···················· | ···················· | ···················· |
| AB | 6811 | hCoV-19/England/205181936/2020\|EPI_ISL_766487\|2020 | ···················· | ···················· | ···················· | ···················· | ···················· | ···················· |
| AB | 6812 | hCoV-19/England/205181938/2020\|EPI_ISL_766488\|2020 | ···················· | ···················· | ···················· | ···················· | ···················· | ···················· |
| AB | 6813 | hCoV-19/England/205181939/2020\|EPI_ISL_766489\|2020 | ···················· | ···················· | ···················· | ···················· | ···················· | ···················· |
| AB | 6814 | hCoV-19/Ireland/D-NVRL-MPX31869/2020\|EPI_ISL_76772 | ···················· | ···················· | ···················· | ···················· | ···················· | ···················· |
| AB | 6815 | hCoV-19/Ireland/CN-NVRL-MPB40005/2020\|EPI_ISL_7677 | ···················· | ···················· | ···················· | ···················· | ···················· | ···················· |
| AB | 6816 | hCoV-19/Ireland/MN-NVRL-MPL33845/2020\|EPI_ISL_7677 | ···················· | ···················· | ···················· | ···················· | ···················· | ···················· |
| AB | 6817 | hCoV-19/Ireland/MN-NVRL-MPW22252/2020\|EPI_ISL_7677 | ···················· | ···················· | ···················· | ···················· | ···················· | ···················· |

| | | | | | | | | |
|---|---|---|---|---|---|---|---|---|
| AB | 6818 hCoV-19/Ireland/MN-NVRL-MPH48921/2020\|EPI_ISL_7677 | ···················· | ···················· | ···················· | ···················· | ···················· | ···················· | |
| AB | 6819 hCoV-19/Ireland/MN-NVRL-MPP19637/2020\|EPI_ISL_7677 | ···················· | ···················· | ···················· | ···················· | ···················· | ···················· | |
| AB | 6820 hCoV-19/Ireland/MN-NVRL-MPW22225/2020\|EPI_ISL_7677 | ···················· | ···················· | ···················· | ···················· | ···················· | ···················· | |
| AB | 6821 hCoV-19/Ireland/CN-NVRL-MPZ29827/2020\|EPI_ISL_7677 | ···················· | ···················· | ···················· | ···················· | ···················· | ···················· | |
| AB | 6822 hCoV-19/Ireland/CO-NVRL-MPZ33926/2020\|EPI_ISL_7677 | ···················· | ···················A | ···················· | ···················· | ···················· | ···················· | 1 |
| AB | 6823 hCoV-19/Ireland/CW-NVRL-MPA64650/2020\|EPI_ISL_7677 | ···················· | ···················· | ···················· | ···················· | ···················· | ···················· | |
| AB | 6824 hCoV-19/Ireland/D-NVRL-MPU65189/2020\|EPI_ISL_76773 | ···················· | ···················· | ···················· | ···················· | ···················· | ···················· | |
| AB | 6825 hCoV-19/Ireland/D-NVRL-MPS61941/2020\|EPI_ISL_76774 | ···················· | ···················· | ···················· | ···················· | ···················· | ···················· | |
| AB | 6826 hCoV-19/Ireland/D-NVRL-MPZ70527/2020\|EPI_ISL_76774 | ···················· | ···················· | ···················· | ···················· | ···················· | ···················· | |
| AB | 6827 hCoV-19/Ireland/D-NVRL-MPS81200/2020\|EPI_ISL_76774 | ···················· | ···················· | ···················· | ···················· | ···················· | ···················· | |
| AB | 6828 hCoV-19/Ireland/D-NVRL-MPA74134/2020\|EPI_ISL_76774 | ···················· | ···················· | ···················· | ···················· | ···················· | ···················· | |
| AB | 6829 hCoV-19/Ireland/D-NVRL-MPB54775/2020\|EPI_ISL_76774 | ···················· | ···················· | ···················· | ···················· | ···················· | ···················· | |
| AB | 6830 hCoV-19/Ireland/D-NVRL-MPW26026/2020\|EPI_ISL_76774 | ···················· | ···················· | ···················· | ···················· | ···················· | ···················· | |
| AB | 6831 hCoV-19/Ireland/D-NVRL-MPU78918/2020\|EPI_ISL_76774 | ···················· | ···················· | ···················· | ···················· | ···················· | ···················· | |
| AB | 6832 hCoV-19/Ireland/D-NVRL-MPS28551/2020\|EPI_ISL_76774 | ···················· | ···················· | ···················· | ···················· | ···················· | ···················· | |
| AB | 6833 hCoV-19/Ireland/D-NVRL-MPA66424/2020\|EPI_ISL_76774 | ···················· | ···················· | ···················· | ···················· | ···················· | ···················· | |
| AB | 6834 hCoV-19/Ireland/D-NVRL-MPS91730/2020\|EPI_ISL_76774 | ···················· | ···················· | ···················· | ···················· | ···················· | ···················· | |
| AB | 6835 hCoV-19/Ireland/D-NVRL-MPD55325/2020\|EPI_ISL_76775 | ···················· | ···················· | ···················· | ···················· | ···················· | ···················· | |
| AB | 6837 hCoV-19/Ireland/D-NVRL-MPD82778/2020\|EPI_ISL_76775 | ···················· | ···················· | ···················· | ···················· | ···················· | ···················· | |
| AB | 6838 hCoV-19/Ireland/D-NVRL-MPA83006/2020\|EPI_ISL_76775 | ···················· | ···················· | ···················· | ···················· | ···················· | ···················· | |
| AB | 6839 hCoV-19/Ireland/D-NVRL-MPZ60412/2020\|EPI_ISL_76775 | ···················· | ···················A | ···················· | ···················· | ···················· | ···················· | 1 |
| AB | 6840 hCoV-19/Ireland/D-NVRL-MPW25565/2020\|EPI_ISL_76775 | ···················· | ···················· | ···················· | ···················· | ···················· | ···················· | |
| AB | 6841 hCoV-19/Ireland/D-NVRL-MPG24146/2020\|EPI_ISL_76775 | ···················· | ···················· | ···················· | ···················· | ···················· | ···················· | |
| AB | 6842 hCoV-19/Ireland/D-NVRL-MPG23468/2020\|EPI_ISL_76775 | ···················· | ···················· | ···················· | ···················· | ···················· | ···················· | |
| AB | 6843 hCoV-19/Ireland/D-NVRL-MPU79359/2020\|EPI_ISL_76775 | ···················· | ···················· | ···················· | ···················· | ···················· | ···················· | |
| AB | 6844 hCoV-19/Ireland/D-NVRL-MPV45546/2020\|EPI_ISL_76775 | ···················· | ···················· | ···················· | ···················· | ···················· | ···················· | |
| AB | 6845 hCoV-19/Ireland/D-NVRL-MPL85584/2020\|EPI_ISL_76776 | ···················· | ···················· | ···················· | ···················· | ···················· | ···················· | |
| AB | 6847 hCoV-19/Ireland/D-NVRL-MPS20970/2020\|EPI_ISL_76776 | ···················· | ···················· | ···················· | ···················· | ···················· | ···················· | |
| AB | 6848 hCoV-19/Ireland/D-NVRL-MPW29674/2020\|EPI_ISL_76776 | ···················· | ···················· | ···················· | ···················· | ···················· | ···················· | |
| AB | 6849 hCoV-19/Ireland/D-NVRL-MPT64850/2020\|EPI_ISL_76776 | ···················· | ···················· | ···················· | ···················· | ···················· | ···················· | |
| AB | 6850 hCoV-19/Ireland/D-NVRL-MPV53968/2020\|EPI_ISL_76776 | ···················· | ···················· | ···················· | ···················· | ···················· | ···················· | |
| AB | 6851 hCoV-19/Ireland/D-NVRL-MPB58618/2020\|EPI_ISL_76776 | ···················· | ···················· | ···················· | ···················· | ···················· | ···················· | |
| AB | 6852 hCoV-19/Ireland/D-NVRL-MPW51790/2020\|EPI_ISL_76776 | ···················· | ···················· | ···················· | ···················· | ···················· | ···················· | |
| AB | 6853 hCoV-19/Ireland/D-NVRL-MPC46630/2020\|EPI_ISL_76776 | ···················· | ···················· | ···················· | ···················· | ···················· | ···················· | |
| AB | 6854 hCoV-19/Ireland/D-NVRL-MPS23027/2020\|EPI_ISL_76777 | ···················· | ···················· | ···················· | ···················· | ···················· | ···················· | |
| AB | 6857 hCoV-19/Ireland/D-NVRL-MPL41406/2020\|EPI_ISL_76777 | ···················· | ···················· | ···················· | ···················· | ···················· | ···················· | |
| AB | 6858 hCoV-19/Ireland/D-NVRL-MPW67031/2020\|EPI_ISL_76777 | ···················· | ···················· | ···················· | ···················· | ···················· | ···················· | |
| AB | 6859 hCoV-19/Ireland/D-NVRL-MPU91828/2020\|EPI_ISL_76777 | ···················· | ···················· | ···················· | ···················· | ···················· | ···················· | |
| AB | 6860 hCoV-19/Ireland/D-NVRL-MPS69308/2020\|EPI_ISL_76777 | ···················· | ···················· | ···················· | ···················· | ···················· | ···················· | |
| AB | 6861 hCoV-19/Ireland/D-NVRL-MPZ11628/2020\|EPI_ISL_76777 | ···················· | ···················· | ···················· | ···················· | ···················· | ···················· | |
| AB | 6862 hCoV-19/Ireland/D-NVRL-MPW27183/2020\|EPI_ISL_76777 | ···················· | ···················· | ···················· | ···················· | ···················· | ···················· | |
| AB | 6863 hCoV-19/Ireland/D-NVRL-MPH55349/2020\|EPI_ISL_76777 | ···················· | ···················· | ···················· | ···················· | ···················· | ···················· | |
| AB | 6864 hCoV-19/Ireland/D-NVRL-MPQ95635/2020\|EPI_ISL_76778 | ···················· | ···················· | ···················· | ···················· | ···················· | ···················· | |
| AB | 6865 hCoV-19/Ireland/D-NVRL-MPQ93159/2020\|EPI_ISL_76778 | ···················· | ···················· | ···················· | ···················· | ···················· | ···················· | |
| AB | 6866 hCoV-19/Ireland/D-NVRL-MPL38042/2020\|EPI_ISL_76778 | ···················· | ···················· | ···················· | ···················· | ···················· | ···················· | |
| AB | 6867 hCoV-19/Ireland/D-NVRL-MPW92967/2020\|EPI_ISL_76778 | ···················· | ···················· | ···················· | ···················· | ···················· | ···················· | |
| AB | 6868 hCoV-19/Ireland/D-NVRL-MPS11026/2020\|EPI_ISL_76778 | ···················· | ···················· | ···················· | ···················· | ···················· | ···················· | |
| AB | 6869 hCoV-19/Ireland/D-NVRL-MPW53420/2020\|EPI_ISL_76778 | ···················· | ···················A | ···················· | ···················· | ···················· | ···················· | 1 |
| AB | 6870 hCoV-19/Ireland/D-NVRL-MPU95698/2020\|EPI_ISL_76778 | ···················· | ···················· | ···················· | ···················· | ···················· | ···················· | |
| AB | 6871 hCoV-19/Ireland/D-NVRL-MPZ66737/2020\|EPI_ISL_76778 | ···················· | ···················· | ···················· | ···················· | ···················· | ···················· | |
| AB | 6872 hCoV-19/Ireland/D-NVRL-MPA71880/2020\|EPI_ISL_76778 | ···················· | ···················· | ···················· | ···················· | ···················· | ···················· | |
| AB | 6873 hCoV-19/Ireland/D-NVRL-MPB62435/2020\|EPI_ISL_76778 | ···················· | ···················· | ···················· | ···················· | ···················· | ···················· | |
| AB | 6874 hCoV-19/Ireland/D-NVRL-MPW49450/2020\|EPI_ISL_76779 | ···················· | ···················· | ···················· | ···················· | ···················· | ···················· | |
| AB | 6875 hCoV-19/Ireland/G-NVRL-MPB77121/2020\|EPI_ISL_76779 | ···················· | ···················· | ···················· | ···················· | ···················· | ···················· | |
| AB | 6876 hCoV-19/Ireland/G-NVRL-MPF73373/2020\|EPI_ISL_76779 | ···················· | ···················· | ···················· | ···················· | ···················· | ···················· | |
| AB | 6877 hCoV-19/Ireland/G-NVRL-MPT13402/2020\|EPI_ISL_76779 | ···················· | ···················· | ···················· | ···················· | ···················· | ···················· | |
| AB | 6878 hCoV-19/Ireland/G-NVRL-MPW79025/2020\|EPI_ISL_76779 | ···················· | ···················· | ···················· | ···················· | ···················· | ···················· | |
| AB | 6879 hCoV-19/Ireland/G-NVRL-MPH77761/2020\|EPI_ISL_76779 | ···················· | ···················· | ···················· | ···················· | ···················· | ···················· | |
| AB | 6880 hCoV-19/Ireland/KE-NVRL-MPA78908/2020\|EPI_ISL_7677 | ···················· | ···················· | ···················· | ···················· | ···················· | ···················· | |
| AB | 6881 hCoV-19/Ireland/KE-NVRL-MPA57711/2020\|EPI_ISL_7677 | ···················· | ···················· | ···················· | ···················· | ···················· | ···················· | |
| AB | 6882 hCoV-19/Ireland/KE-NVRL-MPJ74125/2020\|EPI_ISL_7677 | ···················· | ···················· | ···················· | ···················· | ···················· | ···················· | |
| AB | 6883 hCoV-19/Ireland/KE-NVRL-MPJ83814/2020\|EPI_ISL_7677 | ···················· | ···················· | ···················· | ···················· | ···················· | ···················· | |
| AB | 6884 hCoV-19/England/QEUH-C93877/2020\|EPI_ISL_777105\|20 | ···················· | ···················· | ···················· | ···················· | ···················· | ···················· | |
| AB | 6885 hCoV-19/England/ALDP-C8CC84/2020\|EPI_ISL_777110\|20 | ···················· | ···················· | ···················· | ···················· | ···················· | ···················· | |
| AB | 6886 hCoV-19/England/QEUH-C9383B/2020\|EPI_ISL_777112\|20 | ···················· | ···················· | ···················· | ···················· | ···················· | ···················· | |
| AB | 6887 hCoV-19/England/ALDP-C8C5C5/2020\|EPI_ISL_777113\|20 | ···················· | ···················· | ···················· | ···················· | ···················· | ···················· | |
| AB | 6888 hCoV-19/England/ALDP-C8C194/2020\|EPI_ISL_777119\|20 | ···················· | ···················· | ···················· | ···················· | ···················· | ···················· | |
| AB | 6889 hCoV-19/England/ALDP-C8D11B/2020\|EPI_ISL_777125\|20 | ···················· | ···················· | ···················· | ···················· | ······T············· | ···················· | |
| AB | 6890 hCoV-19/England/ALDP-C8C54D/2020\|EPI_ISL_777126\|20 | ···················· | ···················· | ···················· | ···················· | ···················· | ···················· | |
| AB | 6891 hCoV-19/England/QEUH-C93929/2020\|EPI_ISL_777127\|20 | ···················· | ···················· | ···················· | ···················· | ···················· | ···················· | |
| AB | 6892 hCoV-19/England/ALDP-C8FA69/2020\|EPI_ISL_777128\|20 | ···················· | ···················· | ···················· | ···················· | ···················· | ···················· | |
| AB | 6893 hCoV-19/England/CAMC-C9224D/2020\|EPI_ISL_777133\|20 | ···················· | ···················· | ···················· | ···················· | ···················· | ···················· | |
| AB | 6894 hCoV-19/England/MILK-C6F4F8/2020\|EPI_ISL_778406\|20 | ···················· | ···················· | ···················· | ···················· | ···················· | ···················· | |
| AB | 6895 hCoV-19/England/MILK-C6CC3F/2020\|EPI_ISL_778418\|20 | ···················· | ·A·················· | ···················· | ···················· | ···················· | ···A················ | |
| AB | 6896 hCoV-19/England/QEUH-C645B4/2020\|EPI_ISL_778421\|20 | ···················· | ···················· | ···················· | ···················· | ···················· | ···················· | |
| AB | 6897 hCoV-19/England/QEUH-C6448A/2020\|EPI_ISL_778423\|20 | ···················· | ···················· | ···················· | ···················· | ···················· | ···················· | |
| AB | 6898 hCoV-19/England/MILK-C6CC02/2020\|EPI_ISL_778425\|20 | ···················· | ···················· | ···················· | ···················· | ···················· | ···················· | |
| AB | 6899 hCoV-19/England/MILK-C6F786/2020\|EPI_ISL_778429\|20 | ···················· | ···················· | ···················· | ···················· | ···················· | ···················· | |
| AB | 6900 hCoV-19/England/ALDP-C6A17E/2020\|EPI_ISL_778432\|20 | ···················· | ···················· | ···················· | ···················· | ···················· | ···················· | |
| AB | 6901 hCoV-19/England/QEUH-C6543E/2020\|EPI_ISL_778433\|20 | ···················· | ···················· | ···················· | ···················· | ···················· | ···················· | |

| | | | | | | | | |
|---|---|---|---|---|---|---|---|---|
| AB | 6902 | hCoV-19/England/MILK-C6F892/2020\|EPI_ISL_778438\|20 | ···················· | ···················· | ····················· | ···················· | ···················· | ···················· |
| AB | 6904 | hCoV-19/England/QEUH-C66066/2020\|EPI_ISL_778447\|20 | ···················· | ···················· | ····················· | ···················· | ···················· | ···················· |
| AB | 6905 | hCoV-19/England/QEUH-C6545C/2020\|EPI_ISL_778450\|20 | ···················· | ···················· | ····················· | ···················· | ···················· | ···················· |
| AB | 6906 | hCoV-19/England/MILK-C6F829/2020\|EPI_ISL_778452\|20 | ···················· | ···················· | ····················· | ···················· | ···················· | ···················· |
| AB | 6907 | hCoV-19/England/ALDP-C6B869/2020\|EPI_ISL_778454\|20 | ···················· | ·A·················· | ····················· | ···················· | ···················· | ···A················ |
| AB | 6908 | hCoV-19/England/ALDP-C6BA81/2020\|EPI_ISL_778455\|20 | ···················· | ···················· | ····················· | ···················· | ···················· | ···················· |
| AB | 6909 | hCoV-19/England/MILK-C6F5D7/2020\|EPI_ISL_778458\|20 | ···················· | ···················· | ····················· | ···················· | ···················· | ···················· |
| AB | 6910 | hCoV-19/England/QEUH-C6552C/2020\|EPI_ISL_778459\|20 | ···················· | ···················· | ····················· | ···················· | ···················· | ···················· |
| AB | 6911 | hCoV-19/England/QEUH-C6544D/2020\|EPI_ISL_778463\|20 | ···················· | ···················· | ····················· | ···················· | ···················· | ···················· |
| AB | 6912 | hCoV-19/England/QEUH-C645A5/2020\|EPI_ISL_778465\|20 | ···················· | ···················· | ····················· | ···················· | ···················· | ···················· |
| AB | 6913 | hCoV-19/England/QEUH-CC7A56/2020\|EPI_ISL_760302\|20 | ···················· | ···················· | ····················· | ···················· | ···················· | ···················· |
| AB | 6914 | hCoV-19/England/QEUH-CC4497/2020\|EPI_ISL_760303\|20 | ···················· | ···················· | ····················· | ···················· | ···················· | ···················· |
| AB | 6915 | hCoV-19/England/ALDP-CBF505/2020\|EPI_ISL_760344\|20 | ···················· | ···················· | ····················· | ···················· | ···················· | ···················· |
| AB | 6916 | hCoV-19/England/ALDP-CC0923/2020\|EPI_ISL_760346\|20 | ···················· | ···················· | ····················· | ···················· | ···················· | ···················· |
| AB | 6917 | hCoV-19/England/QEUH-CC5DD7/2020\|EPI_ISL_760347\|20 | ···················· | ···················· | ····················· | ···················· | ···················· | ···················· |
| AB | 6918 | hCoV-19/England/QEUH-CC75A0/2020\|EPI_ISL_760348\|20 | ···················· | ···················· | ····················· | ···················· | ···················· | ···················· |
| AB | 6919 | hCoV-19/England/QEUH-CC77B9/2020\|EPI_ISL_760351\|20 | ···················· | ···················· | ····················· | ···················· | ···················· | ···················· |
| AB | 6920 | hCoV-19/England/QEUH-CC2314/2020\|EPI_ISL_760356\|20 | ···················· | ···················· | ····················· | ···················· | ···················· | ···················· |
| AB | 6921 | hCoV-19/England/ALDP-CBF4EA/2020\|EPI_ISL_760358\|20 | ···················· | ···················· | ····················· | ···················· | ···················· | ···················· |
| AB | 6922 | hCoV-19/England/ALDP-CC0826/2020\|EPI_ISL_760359\|20 | ···················· | ···················· | ····················· | ···················· | ···················· | ···A················ |
| AB | 6923 | hCoV-19/England/ALDP-CBDFFA/2020\|EPI_ISL_760360\|20 | ···················· | ···················· | ····················· | ···················· | ······T············· | ···················· |
| AB | 6924 | hCoV-19/England/QEUH-CC5B19/2020\|EPI_ISL_760361\|20 | ···················· | ···················· | ····················· | ···················· | ···················· | ···················· |
| AB | 6925 | hCoV-19/England/QEUH-CC7BBD/2020\|EPI_ISL_760366\|20 | ···················· | ···················· | ····················· | ···················· | ···················· | ···················· |
| AB | 6926 | hCoV-19/England/QEUH-CC3E21/2020\|EPI_ISL_760368\|20 | ···················· | ···················· | ····················· | ···················· | ···················· | ···················· |
| AB | 6927 | hCoV-19/England/ALDP-CC08EA/2020\|EPI_ISL_760370\|20 | ···················· | ···················· | ····················· | ···················· | ···················· | ···················· |
| AB | 6928 | hCoV-19/England/QEUH-CC3146/2020\|EPI_ISL_760374\|20 | ···················· | ···················· | ····················· | ···················· | ······T············· | ···················· |
| AB | 6929 | hCoV-19/England/QEUH-CC5F86/2020\|EPI_ISL_760375\|20 | ···················· | ···················· | ····················· | ···················· | ···················· | ···················· |
| AB | 6930 | hCoV-19/England/QEUH-CC7B62/2020\|EPI_ISL_760376\|20 | ···················· | ···················· | ····················· | ···················· | ···················· | ···················· |
| AB | 6931 | hCoV-19/England/QEUH-CC2BFE/2020\|EPI_ISL_760379\|20 | ···················· | ···················· | ····················· | ···················· | ···················· | ···················· |
| AB | 6932 | hCoV-19/England/ALDP-CC0905/2020\|EPI_ISL_760386\|20 | ···················· | ·A·················· | ····················· | ···················· | ···················· | ···A················ |
| AB | 6933 | hCoV-19/England/MILK-CA602E/2020\|EPI_ISL_761616\|20 | ···················· | ···················· | ····················· | ···················· | ······T············· | ···················· |
| AB | 6934 | hCoV-19/England/MILK-CA6282/2020\|EPI_ISL_761621\|20 | ···················· | ···················· | ····················· | ···················· | ···················· | ···················· |
| AB | 6935 | hCoV-19/England/MILK-CA66C2/2020\|EPI_ISL_761627\|20 | ···················· | ···················· | ····················· | ···················· | ···················· | ···················· |
| AB | 6936 | hCoV-19/England/MILK-CA7193/2020\|EPI_ISL_761634\|20 | ···················· | ···················· | ····················· | ···················· | ···················· | ···················· |
| AB | 6937 | hCoV-19/England/MILK-CA7254/2020\|EPI_ISL_761636\|20 | ···················· | ···················· | ····················· | ···················· | ···················· | ···················· |
| AB | 6938 | hCoV-19/England/MILK-CA6686/2020\|EPI_ISL_761637\|20 | ···················· | ···················· | ····················· | ···················· | ···················· | ···················· |
| AB | 6939 | hCoV-19/England/MILK-CA68AE/2020\|EPI_ISL_761640\|20 | ···················· | ···················· | ····················· | ···················· | ···················· | ···················· |
| AB | 6940 | hCoV-19/England/ALDP-CB661A/2020\|EPI_ISL_761675\|20 | ···················· | ···················· | ····················· | ···················· | ···················· | ···················· |
| AB | 6941 | hCoV-19/England/ALDP-CB59A9/2020\|EPI_ISL_761676\|20 | ···················· | ···················· | ····················· | ···················· | ···················· | ···················· |
| AB | 6942 | hCoV-19/England/ALDP-CB6AA5/2020\|EPI_ISL_761678\|20 | ···················· | ···················· | ····················· | ···················· | ···················· | ···················· |
| AB | 6943 | hCoV-19/England/ALDP-CB6331/2020\|EPI_ISL_761681\|20 | ···················· | ···················· | ····················· | ···················· | ···················· | ···················· |
| AB | 6944 | hCoV-19/England/ALDP-CB4E50/2020\|EPI_ISL_761682\|20 | ···················· | ···················· | ····················· | ···················· | ···················· | ···················· |
| AB | 6945 | hCoV-19/England/ALDP-CB36FF/2020\|EPI_ISL_761684\|20 | ···················· | ···················· | ····················· | ···················· | ···················· | ···················· |
| AB | 6946 | hCoV-19/England/ALDP-CB5095/2020\|EPI_ISL_761685\|20 | ···················· | ···················· | ····················· | ···················· | ···················· | ···················· |
| AB | 6947 | hCoV-19/England/ALDP-CB60C1/2020\|EPI_ISL_761687\|20 | ···················· | ···················· | ····················· | ···················· | ···················· | ···················· |
| AB | 6948 | hCoV-19/England/ALDP-CB65B3/2020\|EPI_ISL_761689\|20 | ···················· | ···················· | ····················· | ···················· | ···················· | ···················· |
| AB | 6949 | hCoV-19/England/ALDP-CB660B/2020\|EPI_ISL_761690\|20 | ···················· | ···················· | ····················· | ···················· | ···················· | ···················· |
| AB | 6950 | hCoV-19/England/ALDP-CB6AD2/2020\|EPI_ISL_761691\|20 | ···················· | ···················· | ····················· | ···················· | ···················· | ···················· |
| AB | 6951 | hCoV-19/England/ALDP-CB5323/2020\|EPI_ISL_761692\|20 | ···················· | ···················· | ····················· | ···················· | ···················· | ···················· |
| AB | 6952 | hCoV-19/England/ALDP-CB5842/2020\|EPI_ISL_761693\|20 | ···················· | ···················· | ····················· | ···················· | ···················· | ···················· |
| AB | 6953 | hCoV-19/England/ALDP-CB4FD5/2020\|EPI_ISL_761695\|20 | ···················· | ···················· | ····················· | ···················· | ···················· | ···················· |
| AB | 6954 | hCoV-19/England/ALDP-CB4F7B/2020\|EPI_ISL_761697\|20 | ···················· | ·A·················· | ····················· | ···················· | ···················· | ···A················ |
| AB | 6955 | hCoV-19/England/ALDP-CB3C75/2020\|EPI_ISL_761698\|20 | ···················· | ···················· | ····················· | ···················· | ···················· | ···················· |
| AB | 6956 | hCoV-19/England/QEUH-C7F98D/2020\|EPI_ISL_762909\|20 | ···················· | ···················· | ····················· | ···················· | ···················· | ···················· |
| AB | 6957 | hCoV-19/England/MILK-C7A6A9/2020\|EPI_ISL_762914\|20 | ···················· | ···················· | ····················· | ···················· | ···················· | ···················· |
| AB | 6958 | hCoV-19/England/QEUH-C7F97E/2020\|EPI_ISL_762918\|20 | ···················· | ···················· | ····················· | ···················· | ···················· | ···················· |
| AB | 6959 | hCoV-19/England/ALDP-C891C4/2020\|EPI_ISL_762919\|20 | ···················· | ···················· | ····················· | ···················· | ······T············· | ···················· |
| AB | 6960 | hCoV-19/England/QEUH-C7F844/2020\|EPI_ISL_762928\|20 | ···················· | ···················· | ····················· | ···················· | ···················· | ···················· |
| AB | 6961 | hCoV-19/England/ALDP-C8909A/2020\|EPI_ISL_762931\|20 | ···················· | ···················· | ····················· | ···················· | ···················· | ···················· |
| AB | 6962 | hCoV-19/England/QEUH-C7F923/2020\|EPI_ISL_762934\|20 | ···················· | ···················· | ····················· | ···················· | ···················· | ···················· |
| AB | 6963 | hCoV-19/England/MILK-C7A7F1/2020\|EPI_ISL_762945\|20 | ···················· | ···················· | ····················· | ···················· | ······T············· | ···················· |
| AB | 6964 | hCoV-19/England/ALDP-C89276/2020\|EPI_ISL_762951\|20 | ···················· | ···················· | ····················· | ···················· | ···················· | ···················· |
| AB | 6965 | hCoV-19/England/ALDP-C8B2A1/2020\|EPI_ISL_762952\|20 | ···················· | ···················· | ····················· | ···················· | ······T············· | ···················· |
| AB | 6966 | hCoV-19/England/QEUH-C7F835/2020\|EPI_ISL_762956\|20 | ···················· | ···················· | ····················· | ···················· | ···················· | ···················· |
| AB | 6967 | hCoV-19/England/ALDP-C8907C/2020\|EPI_ISL_762957\|20 | ···················· | ···················· | ····················· | ···················· | ···················· | ···················· |
| AB | 6968 | hCoV-19/England/ALDP-C8B21A/2020\|EPI_ISL_762964\|20 | ···················· | ···················· | ····················· | ···················· | ······T············· | ···················· |
| AB | 6969 | hCoV-19/England/QEUH-C7C8B0/2020\|EPI_ISL_762966\|20 | ···················· | ···················· | ····················· | ···················· | ···················· | ···················· |
| AB | 6970 | hCoV-19/Wales/PHWC-4A6BDA/2020\|EPI_ISL_765107\|2020 | ···················· | ···················· | ····················· | ···················· | ···················· | ···················· |
| AB | 6971 | hCoV-19/Wales/PHWC-4A6BE9/2020\|EPI_ISL_765108\|2020 | ···················· | ···················· | ····················· | ···················· | ···················· | ···················· |
| AB | 6972 | hCoV-19/Wales/PHWC-4A6C13/2020\|EPI_ISL_765109\|2020 | ···················· | ···················· | ····················· | ···················· | ···················· | ···················· |
| AB | 6974 | hCoV-19/England/SHEF-10C8FD7/2020\|EPI_ISL_765160\|2 | ···················· | ···················· | ····················· | ···················· | ···················· | ···················· |
| AB | 6975 | hCoV-19/England/SHEF-10C946E/2020\|EPI_ISL_765161\|2 | ···················· | ···················· | ····················· | ···················· | ···················· | ···················· |
| AB | 6976 | hCoV-19/England/SHEF-10C95A7/2020\|EPI_ISL_765162\|2 | ···················· | ···················· | ····················· | ···················· | ···················· | ···················· |
| AB | 6977 | hCoV-19/England/SHEF-10C9B3C/2020\|EPI_ISL_765163\|2 | ···················· | ···················· | ····················· | ···················· | ···················· | ···················· |
| AB | 6978 | hCoV-19/England/SHEF-10C9EF7/2020\|EPI_ISL_765164\|2 | ···················· | ···················· | ····················· | ···················· | ···················· | ···················· |
| AB | 6979 | hCoV-19/England/SHEF-10CA096/2020\|EPI_ISL_765165\|2 | ···················· | ···················· | ····················· | ···················· | ···················· | ···················· |
| AB | 6980 | hCoV-19/England/SHEF-10CA0A5/2020\|EPI_ISL_765166\|2 | ···················· | ···················· | ····················· | ···················· | ······T············· | ···················· |
| AB | 6981 | hCoV-19/England/SHEF-10CA306/2020\|EPI_ISL_765167\|2 | ···················· | ···················· | ····················· | ···················· | ···················· | ···················· |
| AB | 6982 | hCoV-19/England/SHEF-10CC3B9/2020\|EPI_ISL_765168\|2 | ···················· | ···················· | ····················· | ···················· | ···················· | ···················· |
| AB | 6983 | hCoV-19/England/SHEF-10CC52C/2020\|EPI_ISL_765169\|2 | ···················· | ···················· | ····················· | ···················· | ···················· | ···················· |

| | | | | | | | | |
|---|---|---|---|---|---|---|---|---|
| AB | 6984 | hCoV-19/England/SHEF-10CD312/2020\|EPI_ISL_765170\|2 | ···················· | ·A·················· | ···················· | ···················· | ···················· | ···A················ |
| AB | 6985 | hCoV-19/England/SHEF-10CD44C/2020\|EPI_ISL_765171\|2 | ···················· | ···················· | ···················· | ···················· | ···················· | ···················· |
| AB | 6986 | hCoV-19/England/SHEF-10CD813/2020\|EPI_ISL_765172\|2 | ···················· | ···················· | ···················· | ···················· | ···················· | ···················· |
| AB | 6987 | hCoV-19/England/SHEF-10CF77E/2020\|EPI_ISL_765173\|2 | ···················· | ···················· | ···················· | ···················· | ···················· | ···················· |
| AB | 6988 | hCoV-19/England/SHEF-10CFC7F/2020\|EPI_ISL_765174\|2 | ···················· | ···················· | ···················· | ···················· | ···················· | ···················· |
| AB | 6989 | hCoV-19/England/SHEF-10D00F7/2020\|EPI_ISL_765175\|2 | ···················· | ···················· | ···················· | ···················· | ···················· | ···················· |
| AB | 6990 | hCoV-19/England/SHEF-10D02C4/2020\|EPI_ISL_765176\|2 | ···················· | ···················· | ···················· | ···················· | ···················· | ···················· |
| AB | 6991 | hCoV-19/England/SHEF-10D0367/2020\|EPI_ISL_765177\|2 | ···················· | ···················· | ···················· | ···················· | ···················· | ···A················ |
| AB | 6992 | hCoV-19/England/SHEF-10D0394/2020\|EPI_ISL_765178\|2 | ···················· | ···················· | ···················· | ···················· | ······T············· | ···················· |
| AB | 6993 | hCoV-19/England/SHEF-10D0D96/2020\|EPI_ISL_765179\|2 | ···················· | ···················· | ···················· | ···················· | ···················· | ···················· |
| AB | 6994 | hCoV-19/England/SHEF-10D12F0/2020\|EPI_ISL_765180\|2 | ···················· | ···················· | ···················· | ···················· | ······T············· | ···················· |
| AB | 6995 | hCoV-19/England/SHEF-10D14AF/2020\|EPI_ISL_765181\|2 | ···················· | ···················· | ···················· | ···················· | ···················· | ···················· |
| AB | 6996 | hCoV-19/England/SHEF-10D1621/2020\|EPI_ISL_765182\|2 | ···················· | ···················· | ···················· | ···················· | ···················· | ···················· |
| AB | 6997 | hCoV-19/England/SHEF-10D17C4/2020\|EPI_ISL_765183\|2 | ···················· | ···················· | ···················· | ···················· | ···················· | ···················· |
| AB | 6998 | hCoV-19/England/SHEF-10D1894/2020\|EPI_ISL_765184\|2 | ···················· | ···················· | ···················· | ···················· | ···················· | ···················· |
| AB | 6999 | hCoV-19/England/SHEF-10D2620/2020\|EPI_ISL_765185\|2 | ···················· | ···················· | ···················· | ···················· | ···················· | ···················· |
| AB | 7000 | hCoV-19/England/SHEF-10D2857/2020\|EPI_ISL_765186\|2 | ···················· | ···················· | ···················· | ···················· | ···················· | ···················· |
| AB | 7001 | hCoV-19/England/SHEF-10D28C0/2020\|EPI_ISL_765187\|2 | ···················· | ···················· | ···················· | ···················· | ···················· | ···················· |
| AB | 7002 | hCoV-19/England/SHEF-10D3276/2020\|EPI_ISL_765189\|2 | ···················· | ···················· | ···················· | ···················· | ···················· | ···················· |
| AB | 7003 | hCoV-19/England/SHEF-10D35F5/2020\|EPI_ISL_765190\|2 | ···················· | ···················· | ···················· | ···················· | ···················· | ···················· |
| AB | 7004 | hCoV-19/England/SHEF-10D3971/2020\|EPI_ISL_765191\|2 | ···················· | ···················· | ···················· | ···················· | ···················· | ···················· |
| AB | 7005 | hCoV-19/England/SHEF-10D3D84/2020\|EPI_ISL_765192\|2 | ···················· | ···················· | ···················· | ···················· | ···················· | ···················· |
| AB | 7006 | hCoV-19/England/SHEF-10D41A5/2020\|EPI_ISL_765193\|2 | ···················· | ···················· | ···················· | ···················· | ······T············· | ···················· |
| AB | 7007 | hCoV-19/England/SHEF-10D41D2/2020\|EPI_ISL_765194\|2 | ···················· | ···················· | ···················· | ···················· | ······T············· | ···················· |
| AB | 7008 | hCoV-19/England/SHEF-10D43DC/2020\|EPI_ISL_765195\|2 | ···················· | ···················· | ···················· | ···················· | ···················· | ···················· |
| AB | 7009 | hCoV-19/England/SHEF-10D49AD/2020\|EPI_ISL_765196\|2 | ···················· | ···················· | ···················· | ···················· | ···················· | ···················· |
| AB | 7010 | hCoV-19/England/SHEF-10D4BA7/2020\|EPI_ISL_765197\|2 | ···················· | ···················· | ···················· | ···················· | ···················· | ···················· |
| AB | 7011 | hCoV-19/England/SHEF-10D4F9C/2020\|EPI_ISL_765198\|2 | ···················· | ···················· | ···················· | ···················· | ···················· | ···················· |
| AB | 7012 | hCoV-19/England/SHEF-10D55D5/2020\|EPI_ISL_765199\|2 | ···················· | ···················· | ···················· | ···················· | ···················· | ···················· |
| AB | 7013 | hCoV-19/England/205161460/2020\|EPI_ISL_766438\|2020 | ···················· | ···················· | ···················· | ···················· | ···················· | ···················· |
| AB | 7014 | hCoV-19/England/205161461/2020\|EPI_ISL_766439\|2020 | ···················· | ···················· | ···················· | ···················· | ···················· | ···················· |
| AB | 7015 | hCoV-19/England/205161618/2020\|EPI_ISL_766441\|2020 | ···················· | ···················· | ···················· | ···················· | ···················· | ···················· |
| AB | 7016 | hCoV-19/England/205181940/2020\|EPI_ISL_766490\|2020 | ···················· | ···················· | ···················· | ···················· | ···················· | ···················· |
| AB | 7017 | hCoV-19/England/205181941/2020\|EPI_ISL_766491\|2020 | ···················· | ···················· | ···················· | ···················· | ···················· | ···················· |
| AB | 7018 | hCoV-19/England/205181943/2020\|EPI_ISL_766492\|2020 | ···················· | ···················· | ···················· | ···················· | ···················· | ···················· |
| AB | 7019 | hCoV-19/England/205181944/2020\|EPI_ISL_766493\|2020 | ···················· | ···················· | ···················· | ···················· | ···················· | ···················· |
| AB | 7020 | hCoV-19/England/205181945/2020\|EPI_ISL_766494\|2020 | ···················· | ···················· | ···················· | ···················· | ···················· | ···················· |
| AB | 7021 | hCoV-19/England/205181947/2020\|EPI_ISL_766495\|2020 | ···················· | ···················· | ···················· | ···················· | ···················· | ···················· |
| AB | 7022 | hCoV-19/England/205181950/2020\|EPI_ISL_766496\|2020 | ···················· | ···················· | ···················· | ···················· | ···················· | ···················· |
| AB | 7023 | hCoV-19/England/205181951/2020\|EPI_ISL_766497\|2020 | ···················· | ···················· | ···················· | ···················· | ···················· | ···················· |
| AB | 7024 | hCoV-19/England/205181952/2020\|EPI_ISL_766498\|2020 | ···················· | ···················· | ···················· | ···················· | ···················· | ···················· |
| AB | 7025 | hCoV-19/England/205190297/2020\|EPI_ISL_766499\|2020 | ···················· | ···················· | ···················· | ···················· | ···················· | ···················· |
| AB | 7026 | hCoV-19/Ireland/D-NVRL-MPK74461/2020\|EPI_ISL_76776 | ···················· | ···················· | ···················· | ···················· | ···················· | ···················· |
| AB | 7027 | hCoV-19/Luxembourg/LNS7623802/2020\|EPI_ISL_771000\| | ···················· | ···················· | ···················· | ···················· | ···················· | ···················· |
| AB | 7028 | hCoV-19/Luxembourg/LNS2329720/2020\|EPI_ISL_771001\| | ···················· | ···················· | ···················· | ···················· | ···················· | ···················· |
| AB | 7029 | hCoV-19/Luxembourg/LNS4319303/2020\|EPI_ISL_771002\| | ···················· | ···················· | ···················· | ···················· | ···················· | ···················· |
| AB | 7030 | hCoV-19/Luxembourg/LNS9018804/2020\|EPI_ISL_771003\| | ···················· | ···················· | ···················· | ···················· | ···················· | ···················· |
| AB | 7031 | hCoV-19/Luxembourg/LNS6619597/2020\|EPI_ISL_771004\| | ···················· | ···················· | ···················· | ···················· | ···················· | ···················· |
| AB | 7032 | hCoV-19/Luxembourg/LNS3014976/2020\|EPI_ISL_771005\| | ···················· | ···················· | ···················· | ···················· | ···················· | ···················· |
| AB | 7033 | hCoV-19/Luxembourg/LNS5703265/2020\|EPI_ISL_771006\| | ···················· | ···················· | ···················· | ···················· | ···················· | ···················· |
| AB | 7034 | hCoV-19/Luxembourg/LNS8539717/2020\|EPI_ISL_771007\| | ···················· | ···················· | ···················· | ···················· | ···················· | ···················· |
| AB | 7035 | hCoV-19/Luxembourg/LNS2681852/2020\|EPI_ISL_771008\| | ···················· | ···················· | ···················· | ···················· | ···················· | ···················· |
| AB | 7036 | hCoV-19/Luxembourg/LNS3153052/2020\|EPI_ISL_771009\| | ···················· | ···················· | ···················· | ···················· | ···················· | ···················· |
| AB | 7037 | hCoV-19/Luxembourg/LNS4102728/2020\|EPI_ISL_771010\| | ···················· | ···················· | ···················· | ···················· | ···················· | ···················· |
| AB | 7038 | hCoV-19/Luxembourg/LNS5406276/2020\|EPI_ISL_771011\| | ···················· | ···················· | ···················· | ···················· | ···················· | ···················· |
| AB | 7039 | hCoV-19/Luxembourg/LNS4283066/2020\|EPI_ISL_771012\| | ···················· | ···················· | ···················· | ···················· | ···················· | ···················· |
| AB | 7040 | hCoV-19/Luxembourg/LNS7726435/2020\|EPI_ISL_771013\| | ···················· | G··················· | ···················· | ···················· | ···················· | ···················· |
| AB | 7041 | hCoV-19/Luxembourg/LNS4865964/2020\|EPI_ISL_771014\| | ···················· | ···················· | ···················· | ···················· | ······T············· | ···················· |
| AB | 7042 | hCoV-19/Luxembourg/LNS3251574/2020\|EPI_ISL_771015\| | ···················· | ···················· | ···················· | ···················· | ···················· | ···················· |
| AB | 7043 | hCoV-19/Luxembourg/LNS6796846/2020\|EPI_ISL_771016\| | ···················· | ···················· | ···················· | ···················· | ···················· | ···················· |
| AB | 7044 | hCoV-19/Luxembourg/LNS6075331/2020\|EPI_ISL_771017\| | ···················· | ···················· | ···················· | ···················· | ···················· | ···················· |
| AB | 7045 | hCoV-19/Luxembourg/LNS3726428/2020\|EPI_ISL_771018\| | ···················· | ···················· | ···················· | ···················· | ···················· | ···················· |
| AB | 7046 | hCoV-19/Luxembourg/LNS3508944/2020\|EPI_ISL_771019\| | ···················· | ···················· | ···················· | ···················· | ···················· | ···················· |
| AB | 7047 | hCoV-19/Luxembourg/LNS8939103/2020\|EPI_ISL_771020\| | ···················· | ···················· | ···················· | ···················· | ···················· | ···················· |
| AB | 7048 | hCoV-19/Luxembourg/LNS5072399/2020\|EPI_ISL_771021\| | ···················· | ···················· | ···················· | ···················· | ···················· | ···················· |
| AB | 7049 | hCoV-19/Luxembourg/LNS2333525/2020\|EPI_ISL_771022\| | ···················· | ···················· | ···················· | ···················· | ···················· | ···················· |
| AB | 7050 | hCoV-19/Luxembourg/LNS3499878/2020\|EPI_ISL_771023\| | ···················· | ···················· | ···················· | ···················· | ···················· | ···················· |
| AB | 7051 | hCoV-19/Luxembourg/LNS0116765/2020\|EPI_ISL_771024\| | ···················· | ···················· | ···················· | ···················· | ···················· | ···················· |
| AB | 7052 | hCoV-19/Luxembourg/LNS1036385/2020\|EPI_ISL_771025\| | ···················· | ···················· | ···················· | ···················· | ···················· | ···················· |
| AB | 7053 | hCoV-19/Luxembourg/LNS5945877/2020\|EPI_ISL_771026\| | ···················· | ···················· | ···················· | ···················· | ···················· | ···················· |
| AB | 7054 | hCoV-19/Luxembourg/LNS9881878/2020\|EPI_ISL_771027\| | ···················· | ···················· | ···················· | ···················· | ···················· | ···················· |
| AB | 7055 | hCoV-19/Luxembourg/LNS0093188/2020\|EPI_ISL_771028\| | ···················· | ···················· | ···················· | ···················· | ···················· | ···················· |
| AB | 7056 | hCoV-19/Luxembourg/LNS0588798/2020\|EPI_ISL_771029\| | ···················· | ···················· | ···················· | ···················· | ···················· | ···················· |
| AB | 7057 | hCoV-19/Luxembourg/LNS4104090/2020\|EPI_ISL_771030\| | ···················· | ···················· | ···················· | ···················· | ···················· | ···················· |
| AB | 7058 | hCoV-19/Luxembourg/LNS0294350/2020\|EPI_ISL_771031\| | ···················· | ···················· | ···················· | ···················· | ···················· | ···················· |
| AB | 7059 | hCoV-19/Luxembourg/LNS4445166/2020\|EPI_ISL_771032\| | ···················· | ···················· | ···················· | ···················· | ···················· | ···················· |
| AB | 7060 | hCoV-19/Luxembourg/LNS2704357/2020\|EPI_ISL_771033\| | ···················· | ···················· | ···················· | ···················· | ···················· | ···················· |
| AB | 7061 | hCoV-19/Luxembourg/LNS5448126/2020\|EPI_ISL_771034\| | ···················· | ·········A·········· | ···················· | ···················· | ···················· | ···················· |
| AB | 7062 | hCoV-19/Luxembourg/LNS9600772/2020\|EPI_ISL_771035\| | ···················· | ···················· | ···················· | ···················· | ···················· | ···················· |
| AB | 7063 | hCoV-19/Bangladesh/DNAS_DRCIM_isl_1-003/2020\|EPI_I | ···················· | ···················· | ···················· | ···················· | ···················· | ···················· |

AB 7064 hCoV-19/Bangladesh/DNAS_DRCIM_isl_75/2020|EPI_ISL_ ···················· ···················· ···················· ···················· ···················· ····················
AB 7065 hCoV-19/Bangladesh/DNAS_DRCIM_isl_95/2020|EPI_ISL_ ···················· ···················· ···················· ···················· ···················· ····················
AB 7066 hCoV-19/Bangladesh/DNAS_DRCIM_isl_116/2020|EPI_ISL ···················· ···················· ···················· ···················· ···················· ····················
AB 7067 hCoV-19/Bangladesh/DNAS_DRCIM_isl_6299/2020|EPI_IS ···················· ···················· ···················· ···················· ···················· ····················
AB 7068 hCoV-19/Bangladesh/DNAS_DRCIM_isl_6840/2020|EPI_IS ···················· ···················· ···················· ···················· ···················· ····················
AB 7069 hCoV-19/Bangladesh/DNAS_DRCIM_isl_6841/2020|EPI_IS ···················· ···················· ···················· ···················· ···················· ····················
AB 7070 hCoV-19/Bangladesh/DNAS_DRCIM_isl_8225/2020|EPI_IS ···················· ···················· ···················· ···················· ···················· ···············A····
AB 7071 hCoV-19/Bangladesh/DNAS_DRCIM_isl_6300/2020|EPI_IS ···················· ···················· ···················· ···················· ···················· ····················
AB 7072 hCoV-19/Bangladesh/DNAS_DRCIM_isl_2/2020|EPI_ISL_7 ···················· ···················· ···················· ···················· ···················· ····················
AB 7073 hCoV-19/Bangladesh/DNAS_isl_DRCIM_6302/2020|EPI_IS ···················· ···················· ···················· ···················· ···················· ····················
AB 7074 hCoV-19/Bangladesh/DNAS_DRCIM_isl_17/2020|EPI_ISL_ ···················· ···················· ···················· ···················· ···················· ····················
AB 7075 hCoV-19/Bangladesh/DNAS_DRCIM_isl_6301/2020|EPI_IS ···················· ···················· ···················· ···················· ···················· ····················
AB 7076 hCoV-19/Bangladesh/DNAS_DRCIM_isl_4/2020|EPI_ISL_7 ···················· ···················· ···················· ···················· ···················· ····················
AB 7077 hCoV-19/Bangladesh/DNAS_DRCIM_isl_DSIMCH-3/2020|EP ···················· ···················· ···················· ···················· ···················· ····················
AB 7078 hCoV-19/Bangladesh/DNAS_DRCIM_isl_BRB 4/2020|EPI_I ···················· ···················· ···················· ···················· ···················· ····················
AB 7079 hCoV-19/Bangladesh/DNAS_DRCIM_isl_OPS 8/2020|EPI_I ···················· ···················· ···················· ···················· ···················· ····················
AB 7080 hCoV-19/Bangladesh/DNAS_DRCIM_isl_P2-98/2020|EPI_I ···················· ···················· ···················· ···················· ···················· ····················
AB 7081 hCoV-19/Bangladesh/DNAS_DRCIM_isl_Sik-Bip-01/2020| ···················· ···················· ···················· ···················· ···················· ····················
AB 7082 hCoV-19/Bangladesh/DNAS_DRCIM_isl_SM-01/2020|EPI_I ···················· ···················· ···················· ···················· ···················· ····················
AB 7083 hCoV-19/Bangladesh/DNAS_DRCIM_isl_R-88/2020|EPI_IS ···················· ···················· ···················· ···················· ···················· ····················
AB 7084 hCoV-19/Bangladesh/DNAS_isl_P2-104/2020|EPI_ISL_77 ···················· ···················· ···················· ···················· ···················· ····················
AB 7085 hCoV-19/Bangladesh/DNAS_DRCIM_isl_Amal/2020|EPI_IS ···················· ···················· ···················· ···················· ···················· ····················
AB 7086 hCoV-19/Bangladesh/DNAS_DRCIM_isl_MH-03/2020|EPI_I ···················· ···················· ···················· ···················· ···················· ····················
AB 7087 hCoV-19/Bangladesh/DNAS_DRCIM_isl_P2-102/2020|EPI_ ···················· ···················· ···················· ···················· ···················· ····················
AB 7088 hCoV-19/Bangladesh/DNAS_DRCIM_isl_SM-12/2020|EPI_I ···················· ···················· ···················· ···················· ···················· ····················
AB 7089 hCoV-19/Bangladesh/DNAS_DRCIM_isl_MH-02/2020|EPI_I ···················· ···················· ···················· ···················· ···················· ····················
AB 7090 hCoV-19/Bangladesh/DNAS_DRCIM_isl_P2-101/2020|EPI_ ···················· ···················· ···················· ···················· ···················· ····················
AB 7091 hCoV-19/Bangladesh/DNAS_DRCIM_isl_P2-107/2020|EPI_ ···················· ···················· ···················· ···················· ···················· ····················
AB 7092 hCoV-19/Bangladesh/DNAS_DRCIM_isl_Hia/2020|EPI_ISL ···················· ···················· ···················· ···················· ···················· ····················
AB 7093 hCoV-19/Bangladesh/DNAS_DRCIM_isl_P2-106/2020|EPI_ ···················· ···················· ···················· ···················· ···················· ····················
AB 7094 hCoV-19/Bangladesh/DNAS_DRCIM_isl_R-36/2020|EPI_IS ···················· ···················· ···················· ···················· ···················· ····················
AB 7095 hCoV-19/Bangladesh/DNAS_DRCIM_isl_HH_12/2020|EPI_I ···················· ···················· ···················· ···················· ···················· ····················
AB 7096 hCoV-19/Bangladesh/DNAS_DRCIM_isl_KHL-1/2020|EPI_I ···················· ···················· ···················· ···················· ···················· ····················
AB 7097 hCoV-19/Bangladesh/DNAS_DRCIM_isl_B-1/2020|EPI_ISL ···················· ···················· ···················· ···················· ···················· ····················
AB 7098 hCoV-19/Bangladesh/DNAS_DRCIM_isl_Sik-bip 1/2020|E ···················· ···················· ···················· ···················· ···················· ····················
AB 7099 hCoV-19/Bangladesh/DNAS_DRCIM_isl_NUR/2020|EPI_ISL ···················· ···················· ···················· ···················· ···················· ····················
AB 7100 hCoV-19/Bangladesh/DNAS_DRCIM_isl_SIBL-7/2020|EPI_ ···················· ···················· ···················· ···················· ···················· ····················
AB 7101 hCoV-19/Bangladesh/DNAS_DRCIM_isl_SIBL-FH 3/2020|E ···················· ···················· ···················· ···················· ···················· ····················
AB 7102 hCoV-19/Bangladesh/DNAS_DRCIM_isl_IH-04/2020|EPI_I ···················· ···················· ···················· ···················· ···················· ····················
AB 7103 hCoV-19/Bangladesh/DNAS_DRCIM_isl_Hafiz Riad/2020| ···················· ···················· ···················· ···················· ···················· ····················
AB 7104 hCoV-19/Bangladesh/DNAS_DRCIM_isl_SIBL-09/2020|EPI ···················· ···················· ···················· ···················· ···················· ····················
AB 7105 hCoV-19/Bangladesh/DNAS_DRCIM_isl_SIBL-08/2020|EPI ···················· ···················· ···················· ···················· ···················· ····················
AB 7106 hCoV-19/Bangladesh/DNAS_DRCIM_isl_BGH-485/2020|EPI ···················· ···················· ···················· ···················· ···················· ····················
AB 7107 hCoV-19/Bangladesh/DNAS_DRCIM_isl_Tanjima/2020|EPI ···················· ···················· ···················· ···················· ···················· ····················
AB 7108 hCoV-19/Bangladesh/DNAS_DRCIM_isl_DSIMCH-2/2020|EP ···················· ···················· ···················· ···················· ···················· ····················
AB 7109 hCoV-19/Bangladesh/DNAS_DRCIM_isl_SIBL-06/2020|EPI ···················· ···················· ···················· ···················· ···················· ····················
AB 7110 hCoV-19/Bangladesh/DNAS_DRCIM_isl_S-3/2020|EPI_ISL ···················· ···················· ···················· ···················· ···················· ····················
AB 7111 hCoV-19/Bangladesh/DNAS_DRCIM_isl_SIBL-07/2020|EPI ···················· ···················· ···················· ···················· ···················· ····················
AB 7112 hCoV-19/Bangladesh/DNAS_DRCIM_isl_S-5/2020|EPI_ISL ···················· ···················· ···················· ···················· ···················· ····················
AB 7113 hCoV-19/Bangladesh/DNAS_DRICM_isl_Iffat/2020|EPI_I ···················· ···················· ···················· ···················· ···················· ····················
AB 7114 hCoV-19/Bangladesh/DNAS_DRICM_isl_IH-2/2020|EPI_IS ···················· ···················· ···················· ···················· ···················· ····················
AB 7115 hCoV-19/Bangladesh/DNAS_DRICM_isl_BRB-13/2020|EPI_ ···················· ···················· ···················· ···················· ···················· ····················
AB 7116 hCoV-19/Bangladesh/DNAS_DRICM_isl_18927/2020|EPI_I ···················· ···················· ···················· ···················· ···················· ····················
AB 7117 hCoV-19/Bangladesh/DNAS_DRICM_isl_HH-08/2020|EPI_I ···················· ···················· ···················· ···················· ···················· ····················
AB 7118 hCoV-19/Bangladesh/DNAS_DRICM_isl_MM-50/2020|EPI_I ···················· ···················· ···················· ···················· ···················· ····················
AB 7119 hCoV-19/Bangladesh/DNAS_DRICM_isl_R-136/2020|EPI_I ···················· ···················· ···················· ···················· ···················· ····················
AB 7120 hCoV-19/Bangladesh/DNAS_DRICM_isl_MLS-4/2020|EPI_I ···················· ···················· ···················· ···················· ···················· ····················
AB 7121 hCoV-19/Bangladesh/DNAS_DRICM_isl_R-138/2020|EPI_I ···················· ·A·················· ···················· ···················· ···················· ··A·················
AB 7122 hCoV-19/Bangladesh/DNAS_DRICM_isl_EK-91/2020|EPI_I ···················· ·A·················· ···················· ···················· ···················· ··A·················
AB 7123 hCoV-19/Bangladesh/DNAS_DRICM_isl_Enayet/2020|EPI_ ···················· ·A·················· ···················· ···················· ···················· ··A·················
AB 7124 hCoV-19/Bangladesh/DNAS_DRICM_isl_S-02/2020|EPI_IS ···················· ···················· ···················· ···················· ···················· ····················
AB 7125 hCoV-19/Bangladesh/DNAS_DRICM_isl_OPS_3/2020|EPI_I ···················· ·A·················· ···················· ···················· ···················· ··A·················
AB 7126 hCoV-19/Bangladesh/DNAS_DRICM_isl_Ismail/2020|EPI_ ···················· ···················· ···················· ···················· ···················· ····················
AB 7127 hCoV-19/Bangladesh/DNAS_DRICM_isl_MH-13/2020|EPI_I ···················· ···················· ···················· ···················· ···················· ····················
AB 7128 hCoV-19/England/QEUH-C8F489/2020|EPI_ISL_777142|20 ···················· ···················· ···················· ···················· ···················· ····················
AB 7129 hCoV-19/England/ALDP-C8EA6A/2020|EPI_ISL_777144|20 ···················· ···················· ···················· ···················· ···················· ····················
AB 7130 hCoV-19/England/MILK-C90ACF/2020|EPI_ISL_777145|20 ···················· ···················· ···················· ···················· ···················· ····················
AB 7131 hCoV-19/England/QEUH-C66DF6/2020|EPI_ISL_777148|20 ···················· ···················· ···················· ···················· ···················· ····················
AB 7132 hCoV-19/England/QEUH-C91730/2020|EPI_ISL_777149|20 ···················· ···················· ···················· ···················· ···················· ····················
AB 7133 hCoV-19/England/QEUH-C93A17/2020|EPI_ISL_777152|20 ···················· ···················· ···················· ···················· ···················· ····················
AB 7134 hCoV-19/England/ALDP-C8C55C/2020|EPI_ISL_777153|20 ···················· ···················· ···················· ···················· ···················· ····················
AB 7135 hCoV-19/England/QEUH-C935E9/2020|EPI_ISL_777154|20 ···················· ···················· ···················· ···················· ···················· ····················
AB 7136 hCoV-19/England/ALDP-C90133/2020|EPI_ISL_777155|20 ···················· ···················· ···················· ···················· ···················· ····················
AB 7137 hCoV-19/England/ALDP-C8E842/2020|EPI_ISL_777157|20 ···················· ···················· ···················· ···················· ···················· ····················
AB 7138 hCoV-19/England/MILK-C91299/2020|EPI_ISL_777158|20 ···················· ·········T·········· ···················· ···················· ···················· ····················
AB 7139 hCoV-19/England/QEUH-C93710/2020|EPI_ISL_777161|20 ···················· ···················· ···················· ···················· ···················· ····················
AB 7140 hCoV-19/England/ALDP-C8C3AD/2020|EPI_ISL_777163|20 ···················· ···················· ···················· ···················· ···················· ····················
AB 7141 hCoV-19/England/ALDP-C8CC66/2020|EPI_ISL_777166|20 ···················· ···················· ···················· ···················· ···················· ····················
AB 7142 hCoV-19/England/ALDP-C8EA1F/2020|EPI_ISL_777167|20 ···················· ···················· ···················· ···················· ···················· ····················
AB 7143 hCoV-19/England/ALDP-C8E657/2020|EPI_ISL_777168|20 ···················· ···················· ···················· ···················· ···················· ····················

| | | | | | | | | |
|---|---|---|---|---|---|---|---|---|
| AB | 7144 | hCoV-19/England/ALDP-C8EA5B/2020\|EPI_ISL_777170\|20 | ···················· | ···················· | ····················· | ···················· | ···················· | ···················· |
| AB | 7145 | hCoV-19/England/ALDP-C8BC49/2020\|EPI_ISL_777171\|20 | ···················· | ···················· | ····················· | ···················· | ···················· | ···················· |
| AB | 7146 | hCoV-19/England/CAMC-C927B7/2020\|EPI_ISL_777173\|20 | ···················· | ·A·················· | ····················· | ···················· | ···················· | ···A················ |
| AB | 7147 | hCoV-19/England/ALDP-C8C4D7/2020\|EPI_ISL_777177\|20 | ···················· | ···················· | ····················· | ···················· | ···················· | ···················· |
| AB | 7148 | hCoV-19/England/CAMC-C928F0/2020\|EPI_ISL_777181\|20 | ···················· | ···················· | ····················· | ···················· | ···················· | ···················· |
| AB | 7149 | hCoV-19/England/ALDP-C8CCC0/2020\|EPI_ISL_777182\|20 | ···················· | ···················· | ····················· | ···················· | ···················· | ···················· |
| AB | 7150 | hCoV-19/England/ALDP-C8C46E/2020\|EPI_ISL_777183\|20 | ···················· | ···················· | ····················· | ···················· | ···················· | ···················· |
| AB | 7151 | hCoV-19/England/MILK-C90977/2020\|EPI_ISL_777184\|20 | ···················· | ···················· | ····················· | ···················· | ···················· | ···················· |
| AB | 7152 | hCoV-19/England/MILK-C9241A/2020\|EPI_ISL_777187\|20 | ···················· | ···················· | ····················· | ···················· | ···················· | ···················· |
| AB | 7153 | hCoV-19/England/QEUH-C93B41/2020\|EPI_ISL_777191\|20 | ···················· | ···················· | ····················· | ···················· | ···················· | ···················· |
| AB | 7154 | hCoV-19/England/QEUH-C8FCAF/2020\|EPI_ISL_777192\|20 | ···················· | ···················· | ····················· | ···················· | ···················· | ···················· |
| AB | 7155 | hCoV-19/England/ALDP-C8C49B/2020\|EPI_ISL_777193\|20 | ···················· | ···················· | ····················· | ···················· | ···················· | ···················· |
| AB | 7156 | hCoV-19/England/QEUH-C93992/2020\|EPI_ISL_777196\|20 | ···················· | ···················· | ····················· | ···················· | ···················· | ···················· |
| AB | 7157 | hCoV-19/England/ALDP-C6B9A2/2020\|EPI_ISL_778410\|20 | ···················· | ···················· | ····················· | ···················· | ···················· | ···················· |
| AB | 7158 | hCoV-19/England/ALDP-C6B993/2020\|EPI_ISL_778412\|20 | ···················· | ···················· | ····················· | ···················· | ···················· | ···················· |
| AB | 7159 | hCoV-19/England/MILK-C6CC6C/2020\|EPI_ISL_778414\|20 | ···················· | ···················· | ····················· | ···················· | ···················· | ···················· |
| AB | 7160 | hCoV-19/England/ALDP-C6B27A/2020\|EPI_ISL_778416\|20 | ···················· | ···················· | ····················· | ···················· | ···················· | ···················· |
| AB | 7161 | hCoV-19/England/ALDP-C6BA36/2020\|EPI_ISL_778471\|20 | ···················· | ···················· | ····················· | ···················· | ···················· | ···················· |
| AB | 7162 | hCoV-19/England/MILK-C6CC11/2020\|EPI_ISL_778474\|20 | ···················· | ···················· | ····················· | ···················· | ···················· | ···················· |
| AB | 7163 | hCoV-19/England/MILK-C6CA35/2020\|EPI_ISL_778476\|20 | ···················· | ···················· | ····················· | ···················· | ···················· | ···················· |
| AB | 7164 | hCoV-19/England/ALDP-C6A150/2020\|EPI_ISL_778478\|20 | ···················· | ···················· | ····················· | ···················· | ···················· | ···················· |
| AB | 7165 | hCoV-19/England/QEUH-C6694D/2020\|EPI_ISL_778482\|20 | ···················· | ···················· | ····················· | ···················· | ···················· | ···················· |
| AB | 7166 | hCoV-19/England/ALDP-C6BABE/2020\|EPI_ISL_778488\|20 | ···················· | ···················· | ····················· | ···················· | ···················· | ···················· |
| AB | 7167 | hCoV-19/England/ALDP-C6BADC/2020\|EPI_ISL_778491\|20 | ···················· | ···················· | ····················· | ···················· | ·····T·············· | ···················· |
| AB | 7168 | hCoV-19/England/MILK-C6F4CB/2020\|EPI_ISL_778493\|20 | ···················· | ···················· | ····················· | ···················· | ···················· | ···················· |
| AB | 7169 | hCoV-19/England/QEUH-CC3E12/2020\|EPI_ISL_760308\|20 | ···················· | ···················· | ····················· | ···················· | ···················· | ···················· |
| AB | 7170 | hCoV-19/England/QEUH-CC2FA7/2020\|EPI_ISL_760309\|20 | ···················· | ···················· | ····················· | ···················· | ·····T·············· | ···················· |
| AB | 7171 | hCoV-19/England/ALDP-CBE050/2020\|EPI_ISL_760311\|20 | ···················· | ···················· | ····················· | ···················· | ···················· | ···················· |
| AB | 7172 | hCoV-19/England/QEUH-CC26C0/2020\|EPI_ISL_760314\|20 | ···················· | ···················· | ····················· | ···················· | ···················· | ···················· |
| AB | 7173 | hCoV-19/England/QEUH-CC2305/2020\|EPI_ISL_760316\|20 | ···················· | ···················· | ····················· | ···················· | ···················· | ···················· |
| AB | 7174 | hCoV-19/England/QEUH-CC3173/2020\|EPI_ISL_760322\|20 | ···················· | ···················· | ····················· | ···················· | ···················· | ···················· |
| AB | 7175 | hCoV-19/England/QEUH-CC2B85/2020\|EPI_ISL_760323\|20 | ···················· | ···················· | ····················· | ···················· | ···················· | ···················· |
| AB | 7176 | hCoV-19/England/ALDP-CBDD2D/2020\|EPI_ISL_760326\|20 | ···················· | ···················· | ····················· | ···················· | ···················· | ···················· |
| AB | 7177 | hCoV-19/England/QEUH-CC592E/2020\|EPI_ISL_760328\|20 | ···················· | ···················· | ····················· | ···················· | ···················· | ···················· |
| AB | 7178 | hCoV-19/England/QEUH-CC3F5B/2020\|EPI_ISL_760331\|20 | ···················· | ···················· | ····················· | ···················· | ····T··············· | ···················· |
| AB | 7179 | hCoV-19/England/QEUH-CC7ACF/2020\|EPI_ISL_760332\|20 | ···················· | ···················· | ····················· | ···················· | ·····T·············· | ···················· |
| AB | 7180 | hCoV-19/Wales/ALDP-CBE32A/2020\|EPI_ISL_760333\|2020 | ···················· | ···················· | ····················· | ···················· | ···················· | ···················· |
| AB | 7181 | hCoV-19/England/QEUH-CC5D8C/2020\|EPI_ISL_760336\|20 | ···················· | ···················· | ····················· | ···················· | ···················· | ···················· |
| AB | 7182 | hCoV-19/England/QEUH-CC5D9B/2020\|EPI_ISL_760338\|20 | ···················· | ···················· | ····················· | ···················· | ···················· | ···················· |
| AB | 7183 | hCoV-19/England/QEUH-CC75EC/2020\|EPI_ISL_760339\|20 | ···················· | ···················· | ····················· | ···················· | ···················· | ···················· |
| AB | 7184 | hCoV-19/England/ALDP-CBE21E/2020\|EPI_ISL_760341\|20 | ···················· | ···················· | ····················· | ···················· | ···················· | ···················· |
| AB | 7185 | hCoV-19/England/QEUH-CC7643/2020\|EPI_ISL_760343\|20 | ···················· | ···················· | ····················· | ···················· | ·····T·············· | ···················· |
| AB | 7186 | hCoV-19/England/ALDP-CBDC6C/2020\|EPI_ISL_760392\|20 | ···················· | ···················· | ····················· | ···················· | ···················· | ···················· |
| AB | 7187 | hCoV-19/England/QEUH-CC7B35/2020\|EPI_ISL_760394\|20 | ···················· | ···················· | ····················· | ···················· | ···················· | ···················· |
| AB | 7188 | hCoV-19/England/MILK-CA6826/2020\|EPI_ISL_761655\|20 | ···················· | ···················· | ····················· | ···················· | ···················· | ···················· |
| AB | 7189 | hCoV-19/England/MILK-CA663B/2020\|EPI_ISL_761660\|20 | ···················· | ···················· | ····················· | ···················· | ···················· | ···················· |
| AB | 7190 | hCoV-19/England/MILK-CA7069/2020\|EPI_ISL_761661\|20 | ···················· | ·A·······A·········· | ····················· | ···················· | ···················· | ···················· |
| AB | 7191 | hCoV-19/England/ALDP-CB6304/2020\|EPI_ISL_761665\|20 | ···················· | ···················· | ····················· | ···················· | ···················· | ···················· |
| AB | 7192 | hCoV-19/England/ALDP-CB50B3/2020\|EPI_ISL_761666\|20 | ···················· | ···················· | ····················· | ···················· | ···················· | ···················· |
| AB | 7193 | hCoV-19/England/ALDP-CB610A/2020\|EPI_ISL_761670\|20 | ···················· | ···················· | ····················· | ···················· | ···················· | ···A················ |
| AB | 7194 | hCoV-19/England/ALDP-CB502C/2020\|EPI_ISL_761672\|20 | ···················· | ···················· | ····················· | ···················· | ···················· | ···················· |
| AB | 7195 | hCoV-19/England/ALDP-CB60B2/2020\|EPI_ISL_761674\|20 | ···················· | ···················· | ····················· | ···················· | ···················· | ···················· |
| AB | 7196 | hCoV-19/England/ALDP-C8916A/2020\|EPI_ISL_762974\|20 | ···················· | ···················· | ····················· | ···················· | ···················· | ···················· |
| AB | 7197 | hCoV-19/England/ALDP-C890C7/2020\|EPI_ISL_762975\|20 | ···················· | ···················· | ····················· | ···················· | ···················· | ···················· |
| AB | 7198 | hCoV-19/England/ALDP-C8B292/2020\|EPI_ISL_762977\|20 | ···················· | ···················· | ····················· | ···················· | ···A················ | ···················· |
| AB | 7199 | hCoV-19/England/QEUH-C7FB4B/2020\|EPI_ISL_762984\|20 | ···················· | ···················· | ····················· | ···················· | ···················· | ···················· |
| AB | 7200 | hCoV-19/England/SHEF-10D6E8E/2020\|EPI_ISL_765202\|2 | ···················· | ···················· | ····················· | ···················· | ···················· | ···················· |
| AB | 7201 | hCoV-19/England/SHEF-10D78AD/2020\|EPI_ISL_765204\|2 | ···················· | ···················· | ····················· | ···················· | ···················· | ···················· |
| AB | 7202 | hCoV-19/England/SHEF-10D7A2F/2020\|EPI_ISL_765205\|2 | ···················· | ···················· | ····················· | ···················· | ···················· | ···················· |
| AB | 7203 | hCoV-19/England/205190473/2020\|EPI_ISL_766500\|2020 | ···················· | ···················· | ····················· | ···················· | ···················· | ···················· |
| AB | 7204 | hCoV-19/England/205190912/2020\|EPI_ISL_766501\|2020 | ···················· | ···················· | ····················· | ···················· | ···················· | ···················· |
| AB | 7205 | hCoV-19/England/205190929/2020\|EPI_ISL_766502\|2020 | ···················· | ···················· | ····················· | ···················· | ···················· | ···················· |
| AB | 7206 | hCoV-19/England/205190937/2020\|EPI_ISL_766503\|2020 | ···················· | ···················· | ····················· | ···················· | ···················· | ···················· |
| AB | 7207 | hCoV-19/England/205191404/2020\|EPI_ISL_766504\|2020 | ···················· | ···················· | ····················· | ···················· | ···················· | ···················· |
| AB | 7208 | hCoV-19/England/205240596/2020\|EPI_ISL_766505\|2020 | ···················· | ·A·················· | ····················· | ···················· | ···················· | ···A················ |
| AB | 7209 | hCoV-19/England/205260241/2020\|EPI_ISL_766506\|2020 | ···················· | ···················· | ····················· | ···················· | ···················· | ···················· |
| AB | 7210 | hCoV-19/England/205260243/2020\|EPI_ISL_766507\|2020 | ···················· | ···················· | ····················· | ···················· | ···················· | ···················· |
| AB | 7211 | hCoV-19/England/205260244/2020\|EPI_ISL_766508\|2020 | ···················· | ···················· | ····················· | ···················· | ···················· | ···················· |
| AB | 7212 | hCoV-19/England/205260245/2020\|EPI_ISL_766509\|2020 | ···················· | ···················· | ····················· | ···················· | ···················· | ···················· |
| AB | 7213 | hCoV-19/England/205260246/2020\|EPI_ISL_766510\|2020 | ···················· | ···················· | ····················· | ···················· | ···················· | ···················· |
| AB | 7214 | hCoV-19/England/205260247/2020\|EPI_ISL_766511\|2020 | ···················· | ···················· | ····················· | ···················· | ···················· | ···················· |
| AB | 7215 | hCoV-19/England/205260306/2020\|EPI_ISL_766512\|2020 | ···················· | ···················· | ····················· | ···················· | ···················· | ···················· |
| AB | 7216 | hCoV-19/England/205260308/2020\|EPI_ISL_766513\|2020 | ···················· | ···················· | ····················· | ···················· | ···················· | ···················· |
| AB | 7217 | hCoV-19/England/205260312/2020\|EPI_ISL_766514\|2020 | ···················· | ···················· | ····················· | ···················· | ···················· | ···················· |
| AB | 7218 | hCoV-19/England/205260314/2020\|EPI_ISL_766515\|2020 | ···················· | ···················· | ····················· | ···················· | ···················· | ···················· |
| AB | 7219 | hCoV-19/England/205260315/2020\|EPI_ISL_766516\|2020 | ···················· | ···················· | ····················· | ···················· | ···················· | ···················· |
| AB | 7220 | hCoV-19/England/205260317/2020\|EPI_ISL_766517\|2020 | ···················· | ···················· | ····················· | ···················· | ···················· | ···················· |
| AB | 7221 | hCoV-19/England/205260320/2020\|EPI_ISL_766518\|2020 | ···················· | ···················· | ····················· | ···················· | ···················· | ···················· |
| AB | 7222 | hCoV-19/England/205260325/2020\|EPI_ISL_766519\|2020 | ···················· | ···················· | ····················· | ···················· | ···················· | ···················· |
| AB | 7223 | hCoV-19/England/205260395/2020\|EPI_ISL_766520\|2020 | ···················· | ···················· | ····················· | ···················· | ···················· | ···················· |

| | | | | | | | | |
|---|---|---|---|---|---|---|---|---|
| AB | 7224 | hCoV-19/England/205260396/2020\|EPI_ISL_766521\|2020 | .................... | .................... | .................... | .................... | .................... | .................... |
| AB | 7225 | hCoV-19/England/205260397/2020\|EPI_ISL_766522\|2020 | .................... | .................... | .................... | .................... | .................... | .................... |
| AB | 7226 | hCoV-19/England/205260400/2020\|EPI_ISL_766523\|2020 | .................... | .................... | .................... | .................... | .................... | .................... |
| AB | 7227 | hCoV-19/England/205260401/2020\|EPI_ISL_766524\|2020 | .................... | .................... | .................... | .................... | .................... | .................... |
| AB | 7228 | hCoV-19/England/205260402/2020\|EPI_ISL_766525\|2020 | .................... | .................... | .................... | .................... | .................... | .................... |
| AB | 7229 | hCoV-19/England/205260414/2020\|EPI_ISL_766534\|2020 | .................... | .................... | .................... | .................... | .................... | .................... |
| AB | 7230 | hCoV-19/England/205260415/2020\|EPI_ISL_766535\|2020 | .................... | .................... | .................... | .................... | .................... | .................... |
| AB | 7231 | hCoV-19/England/205260416/2020\|EPI_ISL_766536\|2020 | .................... | .................... | .................... | .................... | .................... | .................... |
| AB | 7232 | hCoV-19/Italy/VEN-IZSVe-20RS2155-23_VI/2020\|EPI_IS | .................... | .................... | .................... | .................... | .................... | .................... |
| AB | 7233 | hCoV-19/Italy/VEN-IZSVe-20RS2155-22_VI/2020\|EPI_IS | .................... | .................... | .................... | .................... | ......T............. | .................... |
| AB | 7234 | hCoV-19/Italy/VEN-IZSVe-20RS2155-21_VI/2020\|EPI_IS | .................... | .................... | .................... | .................... | .................... | .................... |
| AB | 7235 | hCoV-19/Italy/VEN-IZSVe-20RS2293-1_VI/2020\|EPI_ISL | .................... | .................... | .................... | .................... | .................... | .................... |
| AB | 7236 | hCoV-19/Italy/VEN-IZSVe-20RS2293-3_VI/2020\|EPI_ISL | .................... | .................... | .................... | .................... | .................... | .................... |
| AB | 7237 | hCoV-19/Italy/VEN-IZSVe-20RS2293-4_VI/2020\|EPI_ISL | .................... | .................... | .................... | .................... | .................... | .................... |
| AB | 7238 | hCoV-19/Italy/VEN-IZSVe-20RS2293-5_VI/2020\|EPI_ISL | .................... | .................... | .................... | .................... | .................... | .................... |
| AB | 7239 | hCoV-19/Italy/VEN-IZSVe-20RS2293-6_VI/2020\|EPI_ISL | .................... | .................... | .................... | .................... | .................... | .................... |
| AB | 7240 | hCoV-19/Switzerland/ZH-UZH-IMV139/2020\|EPI_ISL_766 | .................... | .................... | .................... | .................... | .................... | .................... |
| AB | 7241 | hCoV-19/Switzerland/ZH-UZH-IMV140/2020\|EPI_ISL_766 | .................... | .................... | .................... | .................... | .................... | .................... |
| AB | 7242 | hCoV-19/Switzerland/ZH-UZH-IMV141/2020\|EPI_ISL_766 | .................... | .................... | .................... | .................... | .................... | .................... |
| AB | 7243 | hCoV-19/Sweden/20-52894/2020\|EPI_ISL_766592\|2020-0 | .................... | .................... | .................... | .................... | .................... | .................... |
| AB | 7244 | hCoV-19/Sweden/20-52899/2020\|EPI_ISL_766593\|2020-0 | .................... | .................... | .................... | .................... | .................... | .................... |
| AB | 7245 | hCoV-19/Sweden/20-33501/2020\|EPI_ISL_766594\|2020-1 | .................... | .................... | .................... | .................... | .................... | .................... |
| AB | 7246 | hCoV-19/Sweden/20-33505/2020\|EPI_ISL_766595\|2020-1 | .................... | .................... | .................... | .................... | .................... | .................... |
| AB | 7247 | hCoV-19/Sweden/20-33506/2020\|EPI_ISL_766596\|2020-1 | .................... | .................... | .................... | .................... | .................... | .................... |
| AB | 7248 | hCoV-19/Sweden/20-33509/2020\|EPI_ISL_766597\|2020-1 | .................... | .................... | .................... | .................... | .................... | .................... |
| AB | 7249 | hCoV-19/Sweden/20-33510/2020\|EPI_ISL_766598\|2020-1 | .................... | .................... | .................... | .................... | .................... | .................... |
| AB | 7250 | hCoV-19/Sweden/20-33511/2020\|EPI_ISL_766599\|2020-1 | .................... | .................... | .................... | .................... | .................... | .................... |
| AB | 7251 | hCoV-19/Ireland/KE-NVRL-MPQ65182/2020\|EPI_ISL_7678 | .................... | .................... | .................... | .................... | .................... | .................... |
| AB | 7252 | hCoV-19/Ireland/KE-NVRL-MPP48597/2020\|EPI_ISL_7678 | .................... | .................... | .................... | .................... | .................... | .................... |
| AB | 7253 | hCoV-19/Ireland/KE-NVRL-MPJ90761/2020\|EPI_ISL_7678 | .................... | .................... | .................... | .................... | .................... | .................... |
| AB | 7254 | hCoV-19/Ireland/KK-NVRL-MPB59153/2020\|EPI_ISL_7678 | .................... | .................... | .................... | .................... | .................... | .................... |
| AB | 7255 | hCoV-19/Ireland/KK-NVRL-MPQ56853/2020\|EPI_ISL_7678 | .................... | .................... | .................... | .................... | .................... | .................... |
| AB | 7256 | hCoV-19/Ireland/KK-NVRL-MPQ17909/2020\|EPI_ISL_7678 | .................... | .................... | .................... | .................... | .................... | .................... |
| AB | 7257 | hCoV-19/Ireland/LD-NVRL-MPT68180/2020\|EPI_ISL_7678 | .................... | .................... | .................... | .................... | .................... | .................... |
| AB | 7258 | hCoV-19/Ireland/LH-NVRL-MPZ41611/2020\|EPI_ISL_7678 | .................... | .................... | .................... | .................... | .................... | .................... |
| AB | 7259 | hCoV-19/Ireland/LH-NVRL-MPA80449/2020\|EPI_ISL_7678 | .................... | .................... | .................... | .................... | .................... | .................... |
| AB | 7260 | hCoV-19/Ireland/LH-NVRL-MPB54332/2020\|EPI_ISL_7678 | .................... | .................... | .................... | .................... | .................... | .................... |
| AB | 7261 | hCoV-19/Ireland/LK-NVRL-MPT39150/2020\|EPI_ISL_7678 | .................... | .................... | .................... | .................... | .................... | .................... |
| AB | 7262 | hCoV-19/Ireland/LK-NVRL-MPW54586/2020\|EPI_ISL_7678 | .................... | .................... | .................... | .................... | .................... | .................... |
| AB | 7263 | hCoV-19/Ireland/LK-NVRL-MPK27837/2020\|EPI_ISL_7678 | .................... | .................... | .................... | .................... | .................... | .................... |
| AB | 7264 | hCoV-19/Ireland/LK-NVRL-MPT28673/2020\|EPI_ISL_7678 | .................... | .................... | .................... | .................... | .................... | .................... |
| AB | 7265 | hCoV-19/Ireland/LK-NVRL-MPW50478/2020\|EPI_ISL_7678 | .................... | .................... | .................... | .................... | .................... | .................... |
| AB | 7266 | hCoV-19/Ireland/LS-NVRL-MPS29388/2020\|EPI_ISL_7678 | .................... | .................... | .................... | .................... | .................... | .................... |
| AB | 7267 | hCoV-19/Ireland/MH-NVRL-MPK48258/2020\|EPI_ISL_7678 | .................... | .................... | .................... | .................... | .................... | .................... |
| AB | 7268 | hCoV-19/Ireland/MH-NVRL-MPF36363/2020\|EPI_ISL_7678 | .................... | .................... | .................... | .................... | .................... | .................... |
| AB | 7269 | hCoV-19/Ireland/MH-NVRL-MPR81855/2020\|EPI_ISL_7678 | .................... | .................... | .................... | .................... | .................... | .................... |
| AB | 7270 | hCoV-19/Ireland/MH-NVRL-MPE48476/2020\|EPI_ISL_7678 | .................... | .................... | .................... | .................... | .................... | .................... |
| AB | 7271 | hCoV-19/Ireland/MH-NVRL-MPC80176/2020\|EPI_ISL_7678 | .................... | .................... | .................... | .................... | .................... | .................... |
| AB | 7272 | hCoV-19/Ireland/MN-NVRL-MPB47480/2020\|EPI_ISL_7678 | .................... | .................... | .................... | .................... | .................... | .................... |
| AB | 7273 | hCoV-19/Ireland/MN-NVRL-MPL10032/2020\|EPI_ISL_7678 | .................... | .............A...... | .................... | .................... | .................... | .................... |
| AB | 7274 | hCoV-19/Ireland/MN-NVRL-MPH69450/2020\|EPI_ISL_7678 | .................... | .................... | .................... | .................... | .................... | .................... |
| AB | 7275 | hCoV-19/Ireland/MN-NVRL-MPJ70605/2020\|EPI_ISL_7678 | .................... | .................... | .................... | .................... | .................... | .................... |
| AB | 7276 | hCoV-19/Ireland/OY-NVRL-MPS84145/2020\|EPI_ISL_7678 | .................... | .................... | .................... | .................... | .................... | .................... |
| AB | 7277 | hCoV-19/Ireland/OY-NVRL-MPH73865/2020\|EPI_ISL_7678 | .................... | .................... | .................... | .................... | .................... | .................... |
| AB | 7278 | hCoV-19/Ireland/RN-NVRL-MPA48010/2020\|EPI_ISL_7678 | .................... | .................... | .................... | .................... | .................... | .................... |
| AB | 7279 | hCoV-19/Ireland/RN-NVRL-MPY70268/2020\|EPI_ISL_7678 | .................... | .................... | .................... | .................... | .................... | .................... |
| AB | 7280 | hCoV-19/Ireland/RN-NVRL-MPU16441/2020\|EPI_ISL_7678 | .................... | .................... | .................... | .................... | .................... | .................... |
| AB | 7281 | hCoV-19/Ireland/RN-NVRL-MPH39091/2020\|EPI_ISL_7678 | .................... | .................... | .................... | .................... | .................... | .................... |
| AB | 7282 | hCoV-19/Ireland/WD-NVRL-MPB73322/2020\|EPI_ISL_7678 | .................... | .................... | .................... | .................... | .................... | .................... |
| AB | 7283 | hCoV-19/Ireland/WH-NVRL-MPP18459/2020\|EPI_ISL_7678 | .................... | .................... | .................... | .................... | .................... | .................... |
| AB | 7284 | hCoV-19/Ireland/WH-NVRL-MPW49948/2020\|EPI_ISL_7678 | .................... | .................... | .................... | .................... | .................... | .................... |
| AB | 7285 | hCoV-19/Ireland/WH-NVRL-MPP69791/2020\|EPI_ISL_7678 | .................... | .................... | .................... | .................... | .................... | .................... |
| AB | 7286 | hCoV-19/Ireland/WH-NVRL-MPU16953/2020\|EPI_ISL_7678 | .................... | .................... | .................... | .................... | .................... | .................... |
| AB | 7287 | hCoV-19/Ireland/WW-NVRL-MPJ92577/2020\|EPI_ISL_7678 | .................... | .................... | .................... | .................... | .................... | .................... |
| AB | 7288 | hCoV-19/Ireland/WW-NVRL-MPY84026/2020\|EPI_ISL_7678 | .................... | .................... | .................... | .................... | .................... | .................... |
| AB | 7289 | hCoV-19/Ireland/WW-NVRL-MPN63349/2020\|EPI_ISL_7678 | .................... | .................... | .................... | .................... | .................... | .................... |
| AB | 7290 | hCoV-19/Australia/NSW1284/2020\|EPI_ISL_767850\|2020 | .................... | .................... | .................... | .................... | .................... | .................... |
| AB | 7291 | hCoV-19/Australia/NSW1285/2020\|EPI_ISL_767851\|2020 | .................... | .A.................. | .................... | .................... | .................... | .................... |
| AB | 7292 | hCoV-19/Australia/NSW1286/2020\|EPI_ISL_767852\|2020 | .................... | .A.................. | .................... | .................... | .................... | .................... |
| AB | 7293 | hCoV-19/Australia/NSW1287/2020\|EPI_ISL_767853\|2020 | .................... | .................... | .................... | .................... | .................... | .................... |
| AB | 7294 | hCoV-19/Australia/NSW1288/2020\|EPI_ISL_767854\|2020 | .................... | .................... | .................... | .................... | .................... | .................... |
| AB | 7295 | hCoV-19/Australia/NSW1289/2020\|EPI_ISL_767855\|2020 | .................... | .A.................. | .................... | .................... | .................... | .................... |
| AB | 7296 | hCoV-19/Australia/NSW1290/2020\|EPI_ISL_767856\|2020 | .................... | .................... | .................... | .................... | .................... | .................... |
| AB | 7297 | hCoV-19/Australia/NSW1299/2020\|EPI_ISL_767865\|2020 | .................... | .................... | .................... | .................... | .................... | .................... |
| AB | 7298 | hCoV-19/Australia/NSW1300/2020\|EPI_ISL_767866\|2020 | .................... | .A.................. | .................... | .................... | .................... | ...A................ |
| AB | 7299 | hCoV-19/Australia/NSW1301/2020\|EPI_ISL_767867\|2020 | .................... | .................... | .................... | .................... | .................... | .................... |
| AB | 7300 | hCoV-19/Luxembourg/LNS4702094/2020\|EPI_ISL_771036\| | .................... | .................... | .................... | .................... | .................... | .................... |
| AB | 7301 | hCoV-19/Luxembourg/LNS5168052/2020\|EPI_ISL_771037\| | .................... | .................... | .................... | .................... | .................... | .................... |
| AB | 7302 | hCoV-19/Luxembourg/LNS8312536/2020\|EPI_ISL_771038\| | .................... | .................... | .................... | .................... | .................... | .................... |
| AB | 7303 | hCoV-19/Luxembourg/LNS4943218/2020\|EPI_ISL_771039\| | .................... | .................... | .................... | .................... | .................... | .................... |

| | | | | | | | |
|---|---|---|---|---|---|---|---|
| AB | 7304 hCoV-19/Luxembourg/LNS5748903/2020\|EPI_ISL_771040\| | ···················· | ···················· | ···················· | ···················· | ···················· | ···················· |
| AB | 7305 hCoV-19/Luxembourg/LNS8708455/2020\|EPI_ISL_771041\| | ···················· | ···················· | ···················· | ···················· | ···················· | ···················· |
| AB | 7306 hCoV-19/Luxembourg/LNS6027509/2020\|EPI_ISL_771042\| | ···················· | ···················· | ···················· | ···················· | ···················· | ···················· |
| AB | 7307 hCoV-19/Luxembourg/LNS8464361/2020\|EPI_ISL_771043\| | ···················· | ···················· | ···················· | ···················· | ···················· | ···················· |
| AB | 7308 hCoV-19/Luxembourg/LNS5106192/2020\|EPI_ISL_771044\| | ···················· | ···················· | ···················· | ···················· | ···················· | ···················· |
| AB | 7309 hCoV-19/Luxembourg/LNS0916543/2020\|EPI_ISL_771045\| | ···················· | ···················· | ···················· | ···················· | ···················· | ···················· |
| AB | 7310 hCoV-19/Luxembourg/LNS2810600/2020\|EPI_ISL_771046\| | ···················· | ···················· | ···················· | ···················· | ···················· | ···················· |
| AB | 7311 hCoV-19/Luxembourg/LNS6357507/2020\|EPI_ISL_771047\| | ···················· | ···················· | ···················· | ···················· | ···················· | ···················· |
| AB | 7312 hCoV-19/Luxembourg/LNS8111781/2020\|EPI_ISL_771048\| | ···················· | ···················· | ···················· | ···················· | ···················· | ···················· |
| AB | 7313 hCoV-19/Luxembourg/LNS1649463/2020\|EPI_ISL_771049\| | ···················· | ···················· | ···················· | ···················· | ···················· | ···················· |
| AB | 7314 hCoV-19/Luxembourg/LNS2560142/2020\|EPI_ISL_771050\| | ···················· | ···················· | ···················· | ···················· | ···················· | ···················· |
| AB | 7315 hCoV-19/Luxembourg/LNS6120040/2020\|EPI_ISL_771051\| | ···················· | ···················· | ···················· | ···················· | ···················· | ···················· |
| AB | 7316 hCoV-19/Luxembourg/LNS1002964/2020\|EPI_ISL_771052\| | ···················· | ···················· | ···················· | ···················· | ···················· | ···················· |
| AB | 7317 hCoV-19/Luxembourg/LNS2325120/2020\|EPI_ISL_771053\| | ···················· | ····A··············· | ···················· | ···················· | ···················· | ···················· |
| AB | 7318 hCoV-19/Luxembourg/LNS8374303/2020\|EPI_ISL_771054\| | ···················· | ···················· | ···················· | ···················· | ···················· | ···················· |
| AB | 7319 hCoV-19/Luxembourg/LNS4976308/2020\|EPI_ISL_771055\| | ···················· | ···················· | ···················· | ···················· | ···················· | ···················· |
| AB | 7320 hCoV-19/Luxembourg/LNS7426149/2020\|EPI_ISL_771056\| | ···················· | ···················· | ···················· | ···················· | ···················· | ···················· |
| AB | 7321 hCoV-19/Luxembourg/LNS9135862/2020\|EPI_ISL_771057\| | ···················· | ···················· | ···················· | ···················· | ···················· | ···················· |
| AB | 7322 hCoV-19/Luxembourg/LNS7117233/2020\|EPI_ISL_771058\| | ···················· | ···················· | ···················· | ···················· | ···················· | ···················· |
| AB | 7323 hCoV-19/Luxembourg/LNS5524236/2020\|EPI_ISL_771059\| | ···················· | ···················· | ···················· | ···················· | ···················· | ···················· |
| AB | 7324 hCoV-19/Luxembourg/LNS2954433/2020\|EPI_ISL_771060\| | ···················· | G··················· | ···················· | ···················· | ···················· | ···················· |
| AB | 7325 hCoV-19/Luxembourg/LNS0960880/2020\|EPI_ISL_771061\| | ···················· | ···················· | ···················· | ···················· | ···················· | ···················· |
| AB | 7326 hCoV-19/Luxembourg/LNS8858153/2020\|EPI_ISL_771062\| | ···················· | ···················· | ···················· | ···················· | ···················· | ···················· |
| AB | 7327 hCoV-19/Luxembourg/LNS6943232/2020\|EPI_ISL_771063\| | ···················· | ···················· | ···················· | ···················· | ···················· | ···················· |
| AB | 7328 hCoV-19/Luxembourg/LNS7504894/2020\|EPI_ISL_771064\| | ···················· | ···················· | ···················· | ···················· | ···················· | ···················· |
| AB | 7329 hCoV-19/Luxembourg/LNS2400608/2020\|EPI_ISL_771065\| | ···················· | ···················· | ···················· | ···················· | ···················· | ···················· |
| AB | 7330 hCoV-19/Luxembourg/LNS8892930/2020\|EPI_ISL_771066\| | ···················· | ···················· | ···················· | ···················· | ···················· | ···················· |
| AB | 7331 hCoV-19/Luxembourg/LNS8566339/2020\|EPI_ISL_771067\| | ···················· | ···················· | ···················· | ···················· | ···················· | ···················· |
| AB | 7332 hCoV-19/Luxembourg/LNS4584831/2020\|EPI_ISL_771068\| | ···················· | ···················· | ···················· | ···················· | ···················· | ···················· |
| AB | 7333 hCoV-19/Luxembourg/LNS1583156/2020\|EPI_ISL_771069\| | ···················· | ···················· | ···················· | ···················· | ···················· | ···················· |
| AB | 7334 hCoV-19/Luxembourg/LNS5395059/2020\|EPI_ISL_771070\| | ···················· | ···················· | ···················· | ···················· | ···················· | ···················· |
| AB | 7335 hCoV-19/Luxembourg/LNS6883080/2020\|EPI_ISL_771071\| | ···················· | ···················· | ···················· | ···················· | ···················· | ···················· |
| AB | 7336 hCoV-19/Luxembourg/LNS7425671/2020\|EPI_ISL_771072\| | ···················· | ···················· | ···················· | ···················· | ···················· | ···················· |
| AB | 7337 hCoV-19/Luxembourg/LNS2421976/2020\|EPI_ISL_771073\| | ···················· | ···················· | ···················· | ···················· | ···················· | ···················· |
| AB | 7338 hCoV-19/Luxembourg/LNS2094728/2020\|EPI_ISL_771074\| | ···················· | ···················· | ···················· | ···················· | ···················· | ···················· |
| AB | 7339 hCoV-19/Luxembourg/LNS7326735/2020\|EPI_ISL_771075\| | ···················· | ···················· | ···················· | ···················· | ···················· | ···················· |
| AB | 7340 hCoV-19/Luxembourg/LNS8856783/2020\|EPI_ISL_771076\| | ···················· | ···················· | ···················· | ···················· | ···················· | ···················· |
| AB | 7341 hCoV-19/Luxembourg/LNS1708902/2020\|EPI_ISL_771077\| | ···················· | ····A··············· | ···················· | ···················· | ···················· | ···················· |
| AB | 7342 hCoV-19/Luxembourg/LNS2298315/2020\|EPI_ISL_771078\| | ···················· | ···················· | ···················· | ···················· | ···················· | ···················· |
| AB | 7343 hCoV-19/Luxembourg/LNS1156517/2020\|EPI_ISL_771079\| | ···················· | G··················· | ···················· | ···················· | ···················· | ···················· |
| AB | 7344 hCoV-19/Luxembourg/LNS2478365/2020\|EPI_ISL_771080\| | ···················· | ···················· | ···················· | ···················· | ···················· | ···················· |
| AB | 7345 hCoV-19/Luxembourg/LNS8583405/2020\|EPI_ISL_771081\| | ···················· | ···················· | ···················· | ···················· | ···················· | ···················· |
| AB | 7346 hCoV-19/Luxembourg/LNS1110341/2020\|EPI_ISL_771082\| | ···················· | ···················· | ···················· | ···················· | ···················· | ···················· |
| AB | 7347 hCoV-19/Luxembourg/LNS6437419/2020\|EPI_ISL_771083\| | ···················· | ···················· | ···················· | ···················· | ···················· | ···················· |
| AB | 7348 hCoV-19/Luxembourg/LNS7075819/2020\|EPI_ISL_771084\| | ···················· | ···················· | ···················· | ···················· | ···················· | ···················· |
| AB | 7349 hCoV-19/Luxembourg/LNS7985101/2020\|EPI_ISL_771085\| | ···················· | ···················· | ···················· | ···················· | ···················· | ···················· |
| AB | 7350 hCoV-19/Luxembourg/LNS6379298/2020\|EPI_ISL_771086\| | ···················· | ···················· | ···················· | ···················· | ···················· | ···················· |
| AB | 7351 hCoV-19/Luxembourg/LNS8576610/2020\|EPI_ISL_771087\| | ···················· | G··················· | ···················· | ···················· | ···················· | ···················· |
| AB | 7352 hCoV-19/Luxembourg/LNS3188133/2020\|EPI_ISL_771088\| | ···················· | ···················· | ···················· | ···················· | ···················· | ···················· |
| AB | 7353 hCoV-19/Luxembourg/LNS1399281/2020\|EPI_ISL_771089\| | ···················· | ···················· | ···················· | ···················· | ···················· | ···················· |
| AB | 7354 hCoV-19/Luxembourg/LNS7600165/2020\|EPI_ISL_771090\| | ···················· | ···················· | ···················· | ···················· | ···················· | ···················· |
| AB | 7355 hCoV-19/Luxembourg/LNS4096273/2020\|EPI_ISL_771091\| | ···················· | ···················· | ···················· | ···················· | ···················· | ···················· |
| AB | 7356 hCoV-19/Luxembourg/LNS4306573/2020\|EPI_ISL_771092\| | ···················· | ···················· | ···················· | ···················· | ···················· | ···················· |
| AB | 7357 hCoV-19/Luxembourg/LNS9788161/2020\|EPI_ISL_771093\| | ···················· | G··················· | ···················· | ···················· | ···················· | ···················· |
| AB | 7358 hCoV-19/Luxembourg/LNS2104020/2020\|EPI_ISL_771094\| | ···················· | ···················· | ···················· | ···················· | ···················· | ···················· |
| AB | 7359 hCoV-19/Luxembourg/LNS7045208/2020\|EPI_ISL_771095\| | ···················· | ···················· | ···················· | ···················· | ···················· | ···················· |
| AB | 7360 hCoV-19/Luxembourg/LNS6231323/2020\|EPI_ISL_771096\| | ···················· | ···················· | ···················· | ···················· | ···················· | ···················· |
| AB | 7361 hCoV-19/Luxembourg/LNS2740659/2020\|EPI_ISL_771097\| | ···················· | ···················· | ···················· | ···················· | ···················· | ···················· |
| AB | 7362 hCoV-19/Luxembourg/LNS4950487/2020\|EPI_ISL_771098\| | ···················· | ···················· | ···················· | ···················· | ···················· | ···················· |
| AB | 7363 hCoV-19/Luxembourg/LNS5351629/2020\|EPI_ISL_771099\| | ···················· | ···················· | ···················· | ···················· | ···················· | ···················· |
| AB | 7364 hCoV-19/Bangladesh/DNAS_DRCIM_isl_IH-03/2020\|EPI_I | ···················· | ···················· | G··················· | ···················· | ···················· | ···················· |
| AB | 7365 hCoV-19/Bangladesh/DNAS_DRCIM_isl_KHL-0135/2020\|EP | ···················· | ···················· | ···················· | ···················· | ···················· | ···················· |
| AB | 7366 hCoV-19/Bangladesh/DNAS_DRCIM_isl_HH_13/2020\|EPI_I | ···················· | ···················· | ···················· | ·········N·········· | ···················· | ···················· |
| AB | 7369 hCoV-19/Bangladesh/DNAS_DRCIM_isl_ICHRI-2/2020\|EPI | ···················· | ···················· | ···················· | ···················· | ···················· | ···················· |
| AB | 7370 hCoV-19/Bangladesh/DNAS_DRCIM_isl_R-101/2020\|EPI_I | ···················· | ···················· | ···················· | ···················· | ···················· | ···················· |
| AB | 7371 hCoV-19/Bangladesh/DNAS_DRCIM_isl_Sik-bip 2/2020\|E | ···················· | ···················· | ···················· | ···················· | ···················· | ···················· |
| AB | 7372 hCoV-19/Bangladesh/DNAS_DRCIM_isl_HH-12/2020\|EPI_I | ···················· | ···················· | ···················· | ···················· | ···················· | ···················· |
| AB | 7373 hCoV-19/Bangladesh/DNAS_DRCIM_isl_GL-01/2020\|EPI_I | ···················· | ···················· | ···················· | ···················· | ···················· | ···················· |
| AB | 7374 hCoV-19/Bangladesh/DNAS_DRCIM_isl_HH-11/2020\|EPI_I | ···················· | ···················· | ···················· | ···················· | ···················· | ···················· |
| AB | 7375 hCoV-19/Bangladesh/DNAS_DRCIM_isl_BRB-3/2020\|EPI_I | ···················· | ···················· | ···················· | ···················· | ···················· | ···················· |
| AB | 7376 hCoV-19/Bangladesh/DNAS_DRCIM_isl_MMC-01/2020\|EPI_ | ···················· | ·A·················· | ···················· | ···················· | ···················· | ··A················· |
| AB | 7377 hCoV-19/Bangladesh/DNAS_DRCIM_isl_MMC-02/2020\|EPI_ | ···················· | ·A·················· | ···················· | ···················· | ···················· | ··A················· |
| AB | 7378 hCoV-19/Bangladesh/DNAS_DRCIM_isl_UMCH-1/2020\|EPI_ | ···················· | ···················· | ···················· | ···················· | ···················· | ···················· |
| AB | 7379 hCoV-19/Bangladesh/DNAS_DRCIM_isl_R-97/2020\|EPI_IS | ···················· | ···················· | ···················· | ···················· | ···················· | ···················· |
| AB | 7380 hCoV-19/Bangladesh/DNAS_DRICM_isl_19183/2020\|EPI_I | ···················· | ···················· | ···················· | ···················· | ···················· | ···················· |
| AB | 7381 hCoV-19/Bangladesh/DNAS_DRICM_isl_19189/2020\|EPI_I | ···················· | ···················· | ···················· | ···················· | ···················· | ···················· |
| AB | 7382 hCoV-19/Bangladesh/DNAS_DRICM_isl_ICHRI_4/2020\|EPI | ···················· | ···················· | ···················· | ···················· | ···················· | ···················· |
| AB | 7383 hCoV-19/Bangladesh/DNAS_DRICM_isl_19215/2020\|EPI_I | ···················· | ···················· | ···················· | ···················· | ···················· | ···················· |
| AB | 7384 hCoV-19/Bangladesh/DNAS_DRICM_isl_MH-32/2020\|EPI_I | ···················· | ···················· | ···················· | ···················· | ···················· | ···················· |
| AB | 7385 hCoV-19/Bangladesh/DNAS_DRICM_isl_SIBL-5/2020\|EPI_ | ···················· | ···················· | ···················· | ···················· | ···················· | ···················· |

| | | | | | | | | |
|---|---|---|---|---|---|---|---|---|
| AB | 7386 | hCoV-19/Bangladesh/DNAS_DRICM_isl_MH-22/2020\|EPI_I | ···················· | ···················· | ····················· | ···················· | ···················· | ···················· |
| AB | 7387 | hCoV-19/Bangladesh/DNAS_DRICM_isl_MH-23/2020\|EPI_I | ···················· | ···················· | ····················· | ···················· | ···················· | ···················· |
| AB | 7388 | hCoV-19/Bangladesh/DNAS_DRICM_isl_B-3/2020\|EPI_ISL | ···················· | ···················· | ····················· | ···················· | ···················· | ···················· |
| AB | 7389 | hCoV-19/Bangladesh/GRMDRC/2020\|EPI_ISL_774976\|2020 | ···················· | ···················· | ····················· | ···················· | ···················· | ···················· |
| AB | 7390 | hCoV-19/Bangladesh/DNAS_DRICM_isl_19205/2020\|EPI_I | ···················· | ···················· | ····················· | ···················· | ···················· | ···················· |
| AB | 7391 | hCoV-19/Bangladesh/DNAS_DRICM_isl_B-4/2020\|EPI_ISL | ···················· | ···················· | ····················· | ···················· | ···················· | ···················· |
| AB | 7392 | hCoV-19/Bangladesh/DNAS_DRICM_isl_SIBL_12/2020\|EPI | ···················· | ·A·················· | ····················· | ···················· | ···················· | ··A················· |
| AB | 7393 | hCoV-19/Bangladesh/DNAS_DRICM_isl_B-2/2020\|EPI_ISL | ···················· | ···················· | ····················· | ···················· | ···················· | ···················· |
| AB | 7394 | hCoV-19/Bangladesh/DNAS_DRICM_isl_N-3/2020\|EPI_ISL | ···················· | ···················· | ····················· | ···················· | ···················· | ···················· |
| AB | 7395 | hCoV-19/Bangladesh/DNAS_DRICM_isl_MH-09/2020\|EPI_I | ···················· | ···················· | ····················· | ···················· | ···················· | ···················· |
| AB | 7396 | hCoV-19/Bangladesh/DNAS_DRICM_isl_MR-03/2020\|EPI_I | ···················· | ···················· | ····················· | ···················· | ···················· | ···················· |
| AB | 7397 | hCoV-19/Bangladesh/DNAS_DRICM_isl_18941/2020\|EPI_I | ···················· | ···················· | ····················· | ···················· | ···················· | ···················· |
| AB | 7398 | hCoV-19/England/CAMC-C92104/2020\|EPI_ISL_777218\|20 | ···················· | ···················· | ····················· | ···················· | ···················· | ···················· |
| AB | 7399 | hCoV-19/England/QEUH-C937D4/2020\|EPI_ISL_777219\|20 | ···················· | ···················· | ····················· | ···················· | ···················· | ···················· |
| AB | 7400 | hCoV-19/England/QEUH-C93631/2020\|EPI_ISL_777223\|20 | ···················· | ···················· | ····················· | ···················· | ···················· | ···················· |
| AB | 7401 | hCoV-19/England/MILK-C91897/2020\|EPI_ISL_777224\|20 | ···················· | ···················· | ····················· | ···················· | ···················· | ···················· |
| AB | 7402 | hCoV-19/England/ALDP-C8EA2E/2020\|EPI_ISL_777228\|20 | ···················· | ···················· | ····················· | ···················· | ···················· | ···················· |
| AB | 7403 | hCoV-19/England/QEUH-C93965/2020\|EPI_ISL_777235\|20 | ···················· | ·A·················· | ····················· | ···················· | ···················· | ··A················· |
| AB | 7404 | hCoV-19/England/ALDP-C8D6A3/2020\|EPI_ISL_777240\|20 | ···················· | ···················· | ····················· | ···················· | ···················· | ···················· |
| AB | 7405 | hCoV-19/England/QEUH-C9358F/2020\|EPI_ISL_777241\|20 | ···················· | ···················· | ····················· | ···················· | ···················· | ···················· |
| AB | 7406 | hCoV-19/Wales/ALDP-C8C47D/2020\|EPI_ISL_777243\|2020 | ···················· | ···················· | ····················· | ···················· | ···················· | ···················· |
| AB | 7407 | hCoV-19/England/ALDP-C8FF88/2020\|EPI_ISL_777244\|20 | ···················· | ···················· | ····················· | ···················· | ···················· | ···················· |
| AB | 7408 | hCoV-19/England/QEUH-C937A7/2020\|EPI_ISL_777250\|20 | ···················· | ···················· | ····················· | ···················· | ···················· | ···················· |
| AB | 7409 | hCoV-19/England/QEUH-C937B6/2020\|EPI_ISL_777252\|20 | ···················· | ···················· | ····················· | ···················· | ···················· | ···················· |
| AB | 7410 | hCoV-19/England/QEUH-C938B3/2020\|EPI_ISL_777253\|20 | ···················· | ···················· | ····················· | ···················· | ···················· | ···················· |
| AB | 7411 | hCoV-19/England/QEUH-C948EF/2020\|EPI_ISL_777255\|20 | ···················· | ···················· | ····················· | ···················· | ···················· | ···················· |
| AB | 7412 | hCoV-19/England/ALDP-C6B8C3/2020\|EPI_ISL_778517\|20 | ···················· | ·A·················· | ····················· | ···················· | ···················· | ··A················· |
| AB | 7413 | hCoV-19/England/ALDP-C6A19C/2020\|EPI_ISL_778519\|20 | ···················· | ···················· | ····················· | ···················· | ···················· | ···················· |
| AB | 7414 | hCoV-19/England/MILK-C6CEC0/2020\|EPI_ISL_778550\|20 | ···················· | ·A·················· | ····················· | ···················· | ···················· | ··A················· |
| AB | 7415 | hCoV-19/England/ALDP-C6B896/2020\|EPI_ISL_778556\|20 | ···················· | ···················· | ····················· | ···················· | ···················· | ···················· |
| AB | 7416 | hCoV-19/England/QEUH-C643D8/2020\|EPI_ISL_778560\|20 | ···················· | ···················· | ····················· | ···················· | ···················· | ···················· |
| AB | 7417 | hCoV-19/England/MILK-C6F67A/2020\|EPI_ISL_778565\|20 | ···················· | ···················· | ····················· | ···················· | ···················· | ···················· |
| AB | 7418 | hCoV-19/England/ALDP-C6B887/2020\|EPI_ISL_778569\|20 | ···················· | ···················· | ····················· | ···················· | ···················· | ···················· |
| AB | 7419 | hCoV-19/England/MILK-C6F610/2020\|EPI_ISL_778572\|20 | ···················· | ····A··············· | ····················· | ···················· | ···················· | ···················· |
| AB | 7420 | hCoV-19/England/MILK-C6C67D/2020\|EPI_ISL_778576\|20 | ···················· | ···················· | ····················· | ···················· | ···················· | ···················· |
| AB | 7421 | hCoV-19/England/ALDP-C6BA18/2020\|EPI_ISL_778578\|20 | ···················· | ···················· | ····················· | ···················· | ······T············· | ···················· |
| AB | 7422 | hCoV-19/England/MILK-C6F470/2020\|EPI_ISL_778580\|20 | ···················· | ···················· | ····················· | ···················· | ······T············· | ···················· |
| AB | 7423 | hCoV-19/England/MILK-C6C974/2020\|EPI_ISL_778583\|20 | ···················· | ···················· | ····················· | ···················· | ···················· | ···················· |
| AB | 7424 | hCoV-19/England/ALDP-C6BA72/2020\|EPI_ISL_778584\|20 | ···················· | ···················· | ····················· | ···················· | ······T············· | ···················· |
| AB | 7425 | hCoV-19/England/QEUH-CC7AED/2020\|EPI_ISL_760401\|20 | ···················· | ···················· | ····················· | ···················· | ······T············· | ···················· |
| AB | 7426 | hCoV-19/England/QEUH-CC3BC0/2020\|EPI_ISL_760403\|20 | ···················· | ···················· | ····················· | ···················· | ···················· | ···················· |
| AB | 7427 | hCoV-19/England/QEUH-CC7BAE/2020\|EPI_ISL_760410\|20 | ···················· | ···················· | ····················· | ···················· | ···················· | ···················· |
| AB | 7428 | hCoV-19/England/QEUH-CC7C8D/2020\|EPI_ISL_760413\|20 | ···················· | ···················· | ····················· | ···················· | ···················· | ···················· |
| AB | 7429 | hCoV-19/England/ALDP-CBFE64/2020\|EPI_ISL_760418\|20 | ···················· | ···················· | ····················· | ···················· | ···················· | ···················· |
| AB | 7430 | hCoV-19/England/ALDP-CBDCB7/2020\|EPI_ISL_760419\|20 | ···················· | ···················· | ····················· | ···················· | ······T············· | ···················· |
| AB | 7431 | hCoV-19/England/QEUH-CC3BFD/2020\|EPI_ISL_760422\|20 | ···················· | ···················· | ····················· | ···················· | ···················· | ···················· |
| AB | 7432 | hCoV-19/England/QEUH-CC5B73/2020\|EPI_ISL_760423\|20 | ···················· | ···················· | ····················· | ···················· | ···················· | ···················· |
| AB | 7433 | hCoV-19/England/QEUH-CC44C4/2020\|EPI_ISL_760428\|20 | ···················· | ···················· | ····················· | ···················· | ···················· | ···················· |
| AB | 7434 | hCoV-19/England/QEUH-CC5D5F/2020\|EPI_ISL_760429\|20 | ···················· | ···················· | ····················· | ···················· | ···················· | ···················· |
| AB | 7435 | hCoV-19/England/QEUH-CC5320/2020\|EPI_ISL_760431\|20 | ···················· | ···················· | ····················· | ···················· | ···················· | ···················· |
| AB | 7436 | hCoV-19/England/ALDP-CBE014/2020\|EPI_ISL_760434\|20 | ···················· | ···················· | ····················· | ···················· | ···················· | ···················· |
| AB | 7437 | hCoV-19/England/QEUH-CC5F95/2020\|EPI_ISL_760435\|20 | ···················· | ···················· | ····················· | ···················· | ······T············· | ···················· |
| AB | 7438 | hCoV-19/England/ALDP-CBE1C6/2020\|EPI_ISL_760436\|20 | ···················· | ···················· | ····················· | ···················· | ···················· | ···················· |
| AB | 7439 | hCoV-19/England/QEUH-CC5BBF/2020\|EPI_ISL_760437\|20 | ···················· | ···················· | ····················· | ···················· | ···················· | ···················· |
| AB | 7440 | hCoV-19/England/QEUH-CC52C9/2020\|EPI_ISL_760440\|20 | ···················· | ···················· | ····················· | ···················· | ···················· | ···················· |
| AB | 7441 | hCoV-19/England/ALDP-CBE3C0/2020\|EPI_ISL_760441\|20 | ···················· | ···················· | ····················· | ···················· | ···················· | ···················· |
| AB | 7442 | hCoV-19/England/ALDP-CBDCD5/2020\|EPI_ISL_760442\|20 | ···················· | ···················· | ····················· | ···················· | ···················· | ···················· |
| AB | 7443 | hCoV-19/England/QEUH-CC3B84/2020\|EPI_ISL_760465\|20 | ···················· | ···················· | ····················· | ···················· | ···················· | ···················· |
| AB | 7444 | hCoV-19/England/QEUH-CC3119/2020\|EPI_ISL_760466\|20 | ···················· | ···················· | ····················· | ···················· | ···················· | ···················· |
| AB | 7445 | hCoV-19/England/QEUH-CC75BF/2020\|EPI_ISL_760467\|20 | ···················· | ···················· | ····················· | ···················· | ···················· | ···················· |
| AB | 7446 | hCoV-19/England/ALDP-CBDFAF/2020\|EPI_ISL_760471\|20 | ···················· | ···················· | ····················· | ···················· | ···················· | ···················· |
| AB | 7447 | hCoV-19/England/QEUH-CC5900/2020\|EPI_ISL_760472\|20 | ···················· | ···················· | ····················· | ···················· | ···················· | ···················· |
| AB | 7448 | hCoV-19/England/ALDP-CBFE73/2020\|EPI_ISL_760474\|20 | ···················· | ···················· | ····················· | ···················· | ···················· | ···················· |
| AB | 7449 | hCoV-19/England/ALDP-CBE199/2020\|EPI_ISL_760475\|20 | ···················· | ···················· | ····················· | ···················· | ···················· | ···················· |
| AB | 7450 | hCoV-19/England/QEUH-CC595B/2020\|EPI_ISL_760481\|20 | ···················· | ···················· | ····················· | ···················· | ···················· | ···················· |
| AB | 7451 | hCoV-19/England/QEUH-CC2648/2020\|EPI_ISL_760482\|20 | ···················· | ···················· | ····················· | ···················· | ···················· | ···················· |
| AB | 7452 | hCoV-19/England/ALDP-CBE032/2020\|EPI_ISL_760485\|20 | ···················· | ···················· | ····················· | ···················· | ···················· | ···················· |
| AB | 7453 | hCoV-19/England/ALDP-CBDF90/2020\|EPI_ISL_760486\|20 | ···················· | ···················· | ····················· | ···················· | ···················· | ···················· |
| AB | 7454 | hCoV-19/England/QEUH-CC529C/2020\|EPI_ISL_760487\|20 | ···················· | ···················· | ····················· | ···················· | ···················· | ···················· |
| AB | 7455 | hCoV-19/England/QEUH-CC2BC1/2020\|EPI_ISL_760489\|20 | ···················· | ···················· | ····················· | ···················· | ···················· | ···················· |
| AB | 7456 | hCoV-19/England/QEUH-CC75CE/2020\|EPI_ISL_760493\|20 | ···················· | ···················· | ····················· | ···················· | ···················· | ···················· |
| AB | 7457 | hCoV-19/England/QEUH-CC7CD8/2020\|EPI_ISL_760494\|20 | ···················· | ···················· | ····················· | ···················· | ···················· | ···················· |
| AB | 7458 | hCoV-19/England/ALDP-CBDEDF/2020\|EPI_ISL_760498\|20 | ···················· | ···················· | ····················· | ···················· | ···················· | ···················· |
| AB | 7459 | hCoV-19/England/ALDP-CB55E1/2020\|EPI_ISL_761700\|20 | ···················· | ···················· | ····················· | ···················· | ···················· | ···················· |
| AB | 7460 | hCoV-19/England/ALDP-CB3AD5/2020\|EPI_ISL_761701\|20 | ···················· | ···················· | ····················· | ···················· | ···················· | ···················· |
| AB | 7461 | hCoV-19/England/ALDP-CB5DDA/2020\|EPI_ISL_761702\|20 | ···················· | ···················· | ····················· | ···················· | ···················· | ···················· |
| AB | 7462 | hCoV-19/England/ALDP-CB6137/2020\|EPI_ISL_761703\|20 | ···················· | ···················· | ····················· | ···················· | ···················· | ···················· |
| AB | 7463 | hCoV-19/England/ALDP-CB5183/2020\|EPI_ISL_761704\|20 | ···················· | ···················· | ····················· | ···················· | ···················· | ···················· |
| AB | 7464 | hCoV-19/England/ALDP-CB5B94/2020\|EPI_ISL_761705\|20 | ···················· | ···················· | ····················· | ···················· | ···················· | ···················· |
| AB | 7465 | hCoV-19/England/ALDP-CB446D/2020\|EPI_ISL_761707\|20 | ···················· | ···················· | ····················· | ···················· | ···················· | ···················· |

| | | | | | | | |
|---|---|---|---|---|---|---|---|
| AB | 7466 hCoV-19/England/ALDP-CB5569/2020|EPI_ISL_761709|20 | ···················· | ···················· | ···················· | ···················· | ···················· | ···················· |
| AB | 7467 hCoV-19/England/ALDP-CB5824/2020|EPI_ISL_761711|20 | ···················· | ···················· | ···················· | ···················· | ···················· | ···················· |
| AB | 7468 hCoV-19/Wales/ALDP-CB3BA5/2020|EPI_ISL_761712|2020 | ···················· | ···················· | ···················· | ···················· | ···················· | ···················· |
| AB | 7469 hCoV-19/England/ALDP-CB539C/2020|EPI_ISL_761713|20 | ···················· | ···················· | ···················· | ···················· | ···················· | ···················· |
| AB | 7470 hCoV-19/England/ALDP-CB599A/2020|EPI_ISL_761716|20 | ···················· | ···················· | ···················· | ···················· | ···················· | ···················· |
| AB | 7471 hCoV-19/England/ALDP-CB587F/2020|EPI_ISL_761717|20 | ···················· | ···················· | ···················· | ···················· | ···················· | ···················· |
| AB | 7472 hCoV-19/England/ALDP-CB5271/2020|EPI_ISL_761718|20 | ···················· | ···················· | ···················· | ···················· | ···················· | ···················· |
| AB | 7473 hCoV-19/England/ALDP-CB59F4/2020|EPI_ISL_761719|20 | ···················· | ···················· | ···················· | ···················· | ···················· | ···················· |
| AB | 7474 hCoV-19/England/ALDP-CB596D/2020|EPI_ISL_761722|20 | ···················· | ···················· | ···················· | ···················· | ······T············· | ···················· |
| AB | 7475 hCoV-19/England/ALDP-CB62F8/2020|EPI_ISL_761723|20 | ···················· | ···················· | ···················· | ···················· | ···················· | ···················· |
| AB | 7476 hCoV-19/England/ALDP-CB3677/2020|EPI_ISL_761724|20 | ···················· | ···················· | ···················· | ···················· | ···················· | ···················· |
| AB | 7477 hCoV-19/England/ALDP-CB6717/2020|EPI_ISL_761725|20 | ···················· | ···················· | ···················· | ···················· | ···················· | ···················· |
| AB | 7478 hCoV-19/England/ALDP-CB6119/2020|EPI_ISL_761726|20 | ···················· | ···················· | ···················· | ···················· | ···················· | ···················· |
| AB | 7479 hCoV-19/England/ALDP-CB6726/2020|EPI_ISL_761727|20 | ···················· | ···················· | ···················· | ···················· | ···················· | ···················· |
| AB | 7480 hCoV-19/England/ALDP-CB3AE4/2020|EPI_ISL_761731|20 | ···················· | ···················· | ···················· | ···················· | ···················· | ···················· |
| AB | 7481 hCoV-19/England/ALDP-CB597C/2020|EPI_ISL_761736|20 | ···················· | ···················· | ···················· | ···················· | ···················· | ···················· |
| AB | 7482 hCoV-19/England/ALDP-CB637D/2020|EPI_ISL_761737|20 | ···················· | ···················· | ···················· | ···················· | ···················· | ···················· |
| AB | 7483 hCoV-19/England/ALDP-CB51B0/2020|EPI_ISL_761740|20 | ···················· | ···················· | ···················· | ···················· | ···················· | ···················· |
| AB | 7484 hCoV-19/England/ALDP-CB4816/2020|EPI_ISL_761742|20 | ···················· | ···················· | ···················· | ···················· | ···················· | ···················· |
| AB | 7485 hCoV-19/Wales/ALDP-CB5A6A/2020|EPI_ISL_761745|2020 | ···················· | ···················· | ···················· | ···················· | ···················· | ···················· |
| AB | 7486 hCoV-19/England/ALDP-CB44E5/2020|EPI_ISL_761746|20 | ···················· | ···················· | ···················· | ···················· | ···················· | ···················· |
| AB | 7487 hCoV-19/England/ALDP-CB444F/2020|EPI_ISL_761747|20 | ···················· | ···················· | ···················· | ···················· | ···················· | ···················· |
| AB | 7488 hCoV-19/England/ALDP-CB55C3/2020|EPI_ISL_761750|20 | ···················· | ···················· | ···················· | ···················· | ···················· | ···················· |
| AB | 7489 hCoV-19/England/ALDP-CB5235/2020|EPI_ISL_761751|20 | ···················· | ···················· | ···················· | ···················· | ···················· | ···················· |
| AB | 7490 hCoV-19/England/ALDP-CB61FB/2020|EPI_ISL_761756|20 | ···················· | ···················· | ···················· | ···················· | ···················· | ···················· |
| AB | 7491 hCoV-19/England/ALDP-CB5921/2020|EPI_ISL_761758|20 | ···················· | ···················· | ···················· | ···················· | ···················· | ···················· |
| AB | 7492 hCoV-19/England/ALDP-CB6A2D/2020|EPI_ISL_761759|20 | ···················· | ···················· | ···················· | ···················· | ···················· | ···················· |
| AB | 7493 hCoV-19/England/ALDP-CB561B/2020|EPI_ISL_761762|20 | ···················· | ···················· | ···················· | ···················· | ···················· | ···················· |
| AB | 7494 hCoV-19/England/ALDP-CB510B/2020|EPI_ISL_761764|20 | ···················· | ···················· | ···················· | ···················· | ···················· | ···················· |
| AB | 7495 hCoV-19/England/ALDP-CB3A11/2020|EPI_ISL_761766|20 | ···················· | ···················· | ···················· | ···················· | ···················· | ···················· |
| AB | 7496 hCoV-19/England/ALDP-CB63E6/2020|EPI_ISL_761768|20 | ···················· | ···················· | ···················· | ···················· | ···················· | ···················· |
| AB | 7497 hCoV-19/England/ALDP-CB694E/2020|EPI_ISL_761770|20 | ···················· | ···················· | ···················· | ···················· | ···················· | ···················· |
| AB | 7498 hCoV-19/England/ALDP-CB4412/2020|EPI_ISL_761772|20 | ···················· | ···················· | ···················· | ···················· | ···················· | ···················· |
| AB | 7499 hCoV-19/England/ALDP-CB6A96/2020|EPI_ISL_761798|20 | ···················· | ···················· | ···················· | ···················· | ···················· | ···················· |
| AB | 7500 hCoV-19/England/SHEF-10D6BA5/2020|EPI_ISL_765200|2 | ···················· | ···················· | ···················· | ···················· | ···················· | ···················· |
| AB | 7501 hCoV-19/England/SHEF-10D6D90/2020|EPI_ISL_765201|2 | ···················· | ···················· | ···················· | ···················· | ···················· | ···················· |
| AB | 7502 hCoV-19/England/SHEF-10D75F1/2020|EPI_ISL_765203|2 | ···················· | ···················· | ···················· | ···················· | ···················· | ···················· |
| AB | 7503 hCoV-19/England/SHEF-10D7D9F/2020|EPI_ISL_765206|2 | ···················· | ···················· | ···················· | ···················· | ···················· | ···················· |
| AB | 7504 hCoV-19/England/SHEF-10D85C3/2020|EPI_ISL_765207|2 | ···················· | ·A·················· | ···················· | ···················· | ···················· | ···A················ |
| AB | 7505 hCoV-19/England/SHEF-10D8A1F/2020|EPI_ISL_765208|2 | ···················· | ···················· | ···················· | ···················· | ···················· | ···················· |
| AB | 7506 hCoV-19/USA/OH-ODH-SC35013/2020|EPI_ISL_765209|202 | ···················· | ···················· | ···················· | ···················· | ···················· | ···················· |
| AB | 7507 hCoV-19/Portugal/PT1652/2020|EPI_ISL_765210|2020-1 | ···················· | ···················· | ···················· | ···················· | ···················· | ···················· |
| AB | 7508 hCoV-19/Portugal/PT1666/2020|EPI_ISL_765211|2020-1 | ···················· | ···················· | ···················· | ···················· | ···················· | ···················· |
| AB | 7509 hCoV-19/Portugal/PT1752/2020|EPI_ISL_765212|2020-1 | ···················· | ···················· | ···················· | ···················· | ···················· | ···················· |
| AB | 7510 hCoV-19/Portugal/PT1758/2020|EPI_ISL_765213|2020-1 | ···················· | ···················· | ···················· | ···················· | ···················· | ···················· |
| AB | 7511 hCoV-19/Portugal/PT1767/2020|EPI_ISL_765214|2020-1 | ···················· | ···················· | ···················· | ···················· | ···················· | ···················· |
| AB | 7512 hCoV-19/Portugal/PT1820/2020|EPI_ISL_765215|2020-1 | ···················· | ···················· | ···················· | ···················· | ···················· | ···················· |
| AB | 7513 hCoV-19/Portugal/PT1961/2020|EPI_ISL_765217|2020-1 | ···················· | ···················· | ···················· | ···················· | ···················· | ···················· |
| AB | 7514 hCoV-19/Portugal/PT2038/2020|EPI_ISL_765218|2020-1 | ···················· | ···················· | ···················· | ···················· | ···················· | ···················· |
| AB | 7515 hCoV-19/Portugal/PT1563/2020|EPI_ISL_765219|2020-0 | ···················· | ···················· | ···················· | ···················· | ···················· | ···················· |
| AB | 7517 hCoV-19/Portugal/PT1600/2020|EPI_ISL_765221|2020-0 | ···················· | ····T··············· | ···················· | ···················· | ···················· | ···················· |
| AB | 7518 hCoV-19/USA/OH-ODH-SC35020/2020|EPI_ISL_765222|202 | ···················· | ···················· | ···················· | ···················· | ···················· | ···················· |
| AB | 7519 hCoV-19/England/205260404/2020|EPI_ISL_766526|2020 | ···················· | ···················· | ···················· | ···················· | ···················· | ···················· |
| AB | 7520 hCoV-19/England/205260406/2020|EPI_ISL_766527|2020 | ···················· | ···················· | ···················· | ···················· | ···················· | ···················· |
| AB | 7521 hCoV-19/England/205260407/2020|EPI_ISL_766528|2020 | ···················· | ···················· | ···················· | ···················· | ···················· | ···················· |
| AB | 7522 hCoV-19/England/205260408/2020|EPI_ISL_766529|2020 | ···················· | ···················· | ···················· | ···················· | ···················· | ···················· |
| AB | 7523 hCoV-19/England/205260409/2020|EPI_ISL_766530|2020 | ···················· | ···················· | ···················· | ···················· | ···················· | ···················· |
| AB | 7524 hCoV-19/England/205260411/2020|EPI_ISL_766531|2020 | ···················· | ···················· | ···················· | ···················· | ···················· | ···················· |
| AB | 7525 hCoV-19/England/205260412/2020|EPI_ISL_766532|2020 | ···················· | ···················· | ···················· | ···················· | ···················· | ···················· |
| AB | 7526 hCoV-19/England/205260413/2020|EPI_ISL_766533|2020 | ···················· | ···················· | ···················· | ···················· | ···················· | ···················· |
| AB | 7527 hCoV-19/England/205260417/2020|EPI_ISL_766537|2020 | ···················· | ···················· | ···················· | ···················· | ···················· | ···················· |
| AB | 7528 hCoV-19/England/205260418/2020|EPI_ISL_766538|2020 | ···················· | ···················· | ···················· | ···················· | ···················· | ···················· |
| AB | 7529 hCoV-19/England/205260425/2020|EPI_ISL_766539|2020 | ···················· | ···················· | ···················· | ···················· | ···················· | ···················· |
| AB | 7530 hCoV-19/England/205260426/2020|EPI_ISL_766540|2020 | ···················· | ···················· | ···················· | ···················· | ···················· | ···················· |
| AB | 7531 hCoV-19/England/205260427/2020|EPI_ISL_766541|2020 | ···················· | ···················· | ···················· | ···················· | ···················· | ···················· |
| AB | 7532 hCoV-19/England/205260429/2020|EPI_ISL_766542|2020 | ···················· | ···················· | ···················· | ···················· | ···················· | ···················· |
| AB | 7533 hCoV-19/England/205260431/2020|EPI_ISL_766543|2020 | ···················· | ···················· | ···················· | ···················· | ···················· | ···················· |
| AB | 7534 hCoV-19/England/205260432/2020|EPI_ISL_766544|2020 | ···················· | ···················· | ···················· | ···················· | ···················· | ···················· |
| AB | 7535 hCoV-19/England/205260433/2020|EPI_ISL_766545|2020 | ···················· | ···················· | ···················· | ···················· | ···················· | ···················· |
| AB | 7536 hCoV-19/England/205260434/2020|EPI_ISL_766546|2020 | ···················· | ···················· | ···················· | ···················· | ···················· | ···················· |
| AB | 7537 hCoV-19/England/205260435/2020|EPI_ISL_766547|2020 | ···················· | ···················· | ···················· | ···················· | ···················· | ···················· |
| AB | 7538 hCoV-19/England/205260436/2020|EPI_ISL_766548|2020 | ···················· | ···················· | ···················· | ···················· | ···················· | ···················· |
| AB | 7539 hCoV-19/England/205260437/2020|EPI_ISL_766549|2020 | ···················· | ···················· | ···················· | ···················· | ···················· | ···················· |
| AB | 7540 hCoV-19/England/205260438/2020|EPI_ISL_766550|2020 | ···················· | ···················· | ···················· | ···················· | ···················· | ···················· |
| AB | 7541 hCoV-19/England/205260439/2020|EPI_ISL_766551|2020 | ···················· | ···················· | ···················· | ···················· | ···················· | ···················· |
| AB | 7542 hCoV-19/England/205260440/2020|EPI_ISL_766552|2020 | ···················· | ···················· | ···················· | ···················· | ···················· | ···················· |
| AB | 7543 hCoV-19/England/205260441/2020|EPI_ISL_766553|2020 | ···················· | ···················· | ···················· | ···················· | ···················· | ···················· |
| AB | 7544 hCoV-19/England/205260442/2020|EPI_ISL_766554|2020 | ···················· | ···················· | ···················· | ···················· | ···················· | ···················· |
| AB | 7545 hCoV-19/England/205260443/2020|EPI_ISL_766555|2020 | ···················· | ···················· | ···················· | ···················· | ···················· | ···················· |
| AB | 7546 hCoV-19/England/205261873/2020|EPI_ISL_766556|2020 | ···················· | ···················· | ···················· | ···················· | ···················· | ···················· |

| | | | | | | | | |
|---|---|---|---|---|---|---|---|---|
| AB | 7547 | hCoV-19/England/205261874/2020\|EPI_ISL_766557\|2020 | ···················· | ···················· | ···················· | ···················· | ······T············· | ···················· |
| AB | 7548 | hCoV-19/England/205261876/2020\|EPI_ISL_766558\|2020 | ···················· | ···················· | ···················· | ···················· | ···················· | ···················· |
| AB | 7549 | hCoV-19/England/205261877/2020\|EPI_ISL_766559\|2020 | ···················· | ···················· | ···················· | ···················· | ···················· | ···················· |
| AB | 7550 | hCoV-19/England/205261879/2020\|EPI_ISL_766560\|2020 | ···················· | ···················· | ···················· | ···················· | ···················· | ···················· |
| AB | 7551 | hCoV-19/England/205261880/2020\|EPI_ISL_766561\|2020 | ···················· | ···················· | ···················· | ···················· | ···················· | ···················· |
| AB | 7552 | hCoV-19/England/205261881/2020\|EPI_ISL_766562\|2020 | ···················· | ···················· | ···················· | ···················· | ···················· | ···················· |
| AB | 7553 | hCoV-19/England/205261882/2020\|EPI_ISL_766563\|2020 | ···················· | ···················· | ···················· | ···················· | ···················· | ···················· |
| AB | 7554 | hCoV-19/England/205261886/2020\|EPI_ISL_766564\|2020 | ···················· | ···················· | ···················· | ···················· | ···················· | ···················· |
| AB | 7555 | hCoV-19/England/205261887/2020\|EPI_ISL_766565\|2020 | ···················· | ···················· | ···················· | ···················· | ···················· | ···················· |
| AB | 7556 | hCoV-19/England/205261888/2020\|EPI_ISL_766566\|2020 | ···················· | ···················· | ···················· | ···················· | ···················· | ···················· |
| AB | 7557 | hCoV-19/England/205261890/2020\|EPI_ISL_766567\|2020 | ···················· | ···················· | ···················· | ···················· | ···················· | ···················· |
| AB | 7558 | hCoV-19/England/205261950/2020\|EPI_ISL_766568\|2020 | ···················· | ···················· | ···················· | ···················· | ···················· | ···················· |
| AB | 7559 | hCoV-19/Australia/NSW1291/2020\|EPI_ISL_767857\|2020 | ···················· | ···················· | ···················· | ···················· | ···················· | ···················· |
| AB | 7560 | hCoV-19/Australia/NSW1292/2020\|EPI_ISL_767858\|2020 | ···················· | ···················· | ···················· | ···················· | ···················· | ···················· |
| AB | 7561 | hCoV-19/Australia/NSW1293/2020\|EPI_ISL_767859\|2020 | ···················· | ·A·················· | ···················· | ···················· | ···················· | ···A················ |
| AB | 7562 | hCoV-19/Australia/NSW1294/2020\|EPI_ISL_767860\|2020 | ···················· | ·A·················· | ···················· | ···················· | ···················· | ···A················ |
| AB | 7563 | hCoV-19/Australia/NSW1295/2020\|EPI_ISL_767861\|2020 | ···················· | ···················· | ···················· | ···················· | ···················· | ···················· |
| AB | 7564 | hCoV-19/Australia/NSW1296/2020\|EPI_ISL_767862\|2020 | ···················· | ···················· | ···················· | ···················· | ···················· | ···················· |
| AB | 7565 | hCoV-19/Australia/NSW1297/2020\|EPI_ISL_767863\|2020 | ···················· | ···················· | ···················· | ···················· | ···················· | ···················· |
| AB | 7566 | hCoV-19/Australia/NSW1298/2020\|EPI_ISL_767864\|2020 | ···················· | ···················· | ···················· | ···················· | ···················· | ···················· |
| AB | 7567 | hCoV-19/Australia/NSW1302/2020\|EPI_ISL_767868\|2020 | ···················· | ···················· | ···················· | ···················· | ···················· | ···················· |
| AB | 7568 | hCoV-19/Australia/NSW1303/2020\|EPI_ISL_767869\|2020 | ···················· | ···················· | ···················· | ···················· | ···················· | ···················· |
| AB | 7569 | hCoV-19/Australia/NSW1304/2020\|EPI_ISL_767870\|2020 | ···················· | ···················· | ···················· | ···················· | ···················· | ···················· |
| AB | 7570 | hCoV-19/Australia/NSW1305/2020\|EPI_ISL_767871\|2020 | ···················· | ···················· | ···················· | ···················· | ···················· | ···················· |
| AB | 7571 | hCoV-19/Australia/NSW1306/2020\|EPI_ISL_767872\|2020 | ···················· | ···················· | ···················· | ···················· | ···················· | ···················· |
| AB | 7572 | hCoV-19/Australia/NSW1307/2020\|EPI_ISL_767873\|2020 | ···················· | ···················· | ···················· | ···················· | ···················· | ···················· |
| AB | 7573 | hCoV-19/Australia/NSW1308/2020\|EPI_ISL_767874\|2020 | ···················· | ···················· | ···················· | ···················· | ···················· | ···················· |
| AB | 7574 | hCoV-19/Australia/NSW1309/2020\|EPI_ISL_767875\|2020 | ···················· | ···················· | ···················· | ···················· | ···················· | ···················· |
| AB | 7575 | hCoV-19/Australia/NSW1310/2020\|EPI_ISL_767876\|2020 | ···················· | ···················· | ···················· | ···················· | ···················· | ···················· |
| AB | 7576 | hCoV-19/Australia/NSW1311/2020\|EPI_ISL_767877\|2020 | ···················· | ···················· | ···················· | ···················· | ···················· | ···················· |
| AB | 7577 | hCoV-19/Australia/NSW1312/2020\|EPI_ISL_767878\|2020 | ···················· | ···················· | ···················· | ···················· | ···················· | ···················· |
| AB | 7578 | hCoV-19/Australia/NSW1313/2020\|EPI_ISL_767879\|2020 | ···················· | ···················· | ···················· | ···················· | ···················· | ···················· |
| AB | 7579 | hCoV-19/Australia/NSW1314/2020\|EPI_ISL_767880\|2020 | ···················· | ···················· | ···················· | ···················· | ···················· | ···················· |
| AB | 7580 | hCoV-19/Australia/NSW1315/2020\|EPI_ISL_767881\|2020 | ···················· | ···················· | ···················· | ···················· | ···················· | ···················· |
| AB | 7581 | hCoV-19/Australia/NSW1316/2020\|EPI_ISL_767882\|2020 | ···················· | ···················· | ···················· | ···················· | ···················· | ···················· |
| AB | 7582 | hCoV-19/Australia/NSW1317/2020\|EPI_ISL_767883\|2020 | ···················· | ···················· | ···················· | ···················· | ···················· | ···················· |
| AB | 7583 | hCoV-19/Australia/NSW1318/2020\|EPI_ISL_767884\|2020 | ···················· | ···················· | ···················· | ···················· | ···················· | ···················· |
| AB | 7584 | hCoV-19/Australia/NSW1319/2020\|EPI_ISL_767885\|2020 | ···················· | ···················· | ···················· | ···················· | ···················· | ···················· |
| AB | 7585 | hCoV-19/Australia/NSW1320/2020\|EPI_ISL_767886\|2020 | ···················· | ···················· | ···················· | ···················· | ···················· | ···················· |
| AB | 7586 | hCoV-19/Australia/NSW1321/2020\|EPI_ISL_767887\|2020 | ···················· | ···················· | ···················· | ···················· | ···················· | ···················· |
| AB | 7587 | hCoV-19/Australia/NSW1322/2020\|EPI_ISL_767888\|2020 | ···················· | ···················· | ···················· | ···················· | ···················· | ···················· |
| AB | 7588 | hCoV-19/Australia/NSW1323/2020\|EPI_ISL_767889\|2020 | ···················· | ···················· | ···················· | ···················· | ···················· | ···················· |
| AB | 7589 | hCoV-19/Australia/NSW1324/2020\|EPI_ISL_767890\|2020 | ···················· | ···················· | ···················· | ···················· | ···················· | ···················· |
| AB | 7590 | hCoV-19/Australia/NSW1325/2020\|EPI_ISL_767891\|2020 | ···················· | ·A·················· | ···················· | ···················· | ···················· | ···················· |
| AB | 7591 | hCoV-19/Australia/NSW1326/2020\|EPI_ISL_767892\|2020 | ···················· | ···················· | ···················· | ···················· | ···················· | ···················· |
| AB | 7592 | hCoV-19/Australia/NSW1327/2020\|EPI_ISL_767893\|2020 | ···················· | ···················· | ···················· | ···················· | ···················· | ···················· |
| AB | 7593 | hCoV-19/Australia/NSW1328/2020\|EPI_ISL_767894\|2020 | ···················· | ···················· | ···················· | ···················· | ···················· | ···················· |
| AB | 7594 | hCoV-19/Australia/NSW1329/2020\|EPI_ISL_767895\|2020 | ···················· | ···················· | ···················· | ···················· | ···················· | ···················· |
| AB | 7595 | hCoV-19/Australia/NSW1330/2020\|EPI_ISL_767896\|2020 | ···················· | ···················· | ···················· | ···················· | ···················· | ···················· |
| AB | 7596 | hCoV-19/Australia/NSW1331/2020\|EPI_ISL_767897\|2020 | ···················· | ···················· | ···················· | ···················· | ···················· | ···················· |
| AB | 7597 | hCoV-19/Australia/NSW1332/2020\|EPI_ISL_767898\|2020 | ···················· | ···················· | ···················· | ···················· | ···················· | ···················· |
| AB | 7598 | hCoV-19/Australia/NSW1333/2020\|EPI_ISL_767899\|2020 | ···················· | ···················· | ···················· | ···················· | ···················· | ···················· |
| AB | 7599 | hCoV-19/Luxembourg/LNS8080814/2020\|EPI_ISL_771103\| | ···················· | G··················· | ···················· | ···················· | ···················· | ···················· |
| AB | 7600 | hCoV-19/Luxembourg/LNS0486612/2020\|EPI_ISL_771104\| | ···················· | ····A··············· | ···················· | ···················· | ···················· | ···················· |
| AB | 7601 | hCoV-19/Luxembourg/LNS2910621/2020\|EPI_ISL_771105\| | ···················· | G··················· | ···················· | ···················· | ···················· | ···················· |
| AB | 7602 | hCoV-19/Luxembourg/LNS6736919/2020\|EPI_ISL_771113\| | ···················· | ···················· | ···················· | ···················· | ···················· | ···················· |
| AB | 7603 | hCoV-19/Luxembourg/LNS0333025/2020\|EPI_ISL_771114\| | ···················· | ···················· | ···················· | ···················· | ···················· | ···················· |
| AB | 7604 | hCoV-19/Luxembourg/LNS5340325/2020\|EPI_ISL_771115\| | ···················· | ···················· | ···················· | ···················· | ···················· | ···················· |
| AB | 7605 | hCoV-19/Luxembourg/LNS7860492/2020\|EPI_ISL_771116\| | ···················· | ···················· | ···················· | ···················· | ···················· | ···················· |
| AB | 7606 | hCoV-19/Luxembourg/LNS4335636/2020\|EPI_ISL_771117\| | ···················· | ···················· | ···················· | ···················· | ···················· | ···················· |
| AB | 7607 | hCoV-19/Luxembourg/LNS2366276/2020\|EPI_ISL_771118\| | ···················· | ···················· | ···················· | ···················· | ···················· | ···················· |
| AB | 7608 | hCoV-19/Luxembourg/LNS1820740/2020\|EPI_ISL_771119\| | ···················· | ···················· | ···················· | ···················· | ···················· | ···················· |
| AB | 7609 | hCoV-19/Luxembourg/LNS8282542/2020\|EPI_ISL_771120\| | ···················· | ···················· | ···················· | ···················· | ···················· | ···················· |
| AB | 7610 | hCoV-19/Luxembourg/LNS7219474/2020\|EPI_ISL_771121\| | ···················· | ···················· | ···················· | ···················· | ···················· | ···················· |
| AB | 7611 | hCoV-19/Luxembourg/LNS7639610/2020\|EPI_ISL_771122\| | ···················· | G··················· | ···················· | ···················· | ···················· | ···················· |
| AB | 7612 | hCoV-19/Luxembourg/LNS2718564/2020\|EPI_ISL_771123\| | ···················· | ···················· | ···················· | ···················· | ···················· | ···················· |
| AB | 7613 | hCoV-19/Luxembourg/LNS9024343/2020\|EPI_ISL_771124\| | ···················· | G··················· | ···················· | ···················· | ···················· | ···················· |
| AB | 7614 | hCoV-19/Luxembourg/LNS3144077/2020\|EPI_ISL_771125\| | ···················· | ···················· | ···················· | ···················· | ···················· | ···················· |
| AB | 7615 | hCoV-19/Luxembourg/LNS6262482/2020\|EPI_ISL_771126\| | ···················· | ···················· | ···················· | ···················· | ···················· | ···················· |
| AB | 7616 | hCoV-19/Luxembourg/LNS4661498/2020\|EPI_ISL_771127\| | ···················· | ···················· | ···················· | ···················· | ···················· | ···················· |
| AB | 7617 | hCoV-19/Luxembourg/LNS1269461/2020\|EPI_ISL_771128\| | ···················· | ···················· | ···················· | ···················· | ···················· | ···················· |
| AB | 7618 | hCoV-19/Luxembourg/LNS7282970/2020\|EPI_ISL_771129\| | ···················· | ···················· | ···················· | ···················· | ···················· | ···················· |
| AB | 7619 | hCoV-19/Luxembourg/LNS1385707/2020\|EPI_ISL_771130\| | ···················· | ···················· | ···················· | ···················· | ···················· | ···················· |
| AB | 7620 | hCoV-19/Luxembourg/LNS9709367/2020\|EPI_ISL_771131\| | ···················· | G··················· | ···················· | ···················· | ···················· | ···················· |
| AB | 7621 | hCoV-19/Luxembourg/LNS6023003/2020\|EPI_ISL_771132\| | ···················· | G··················· | ···················· | ···················· | ···················· | ···················· |
| AB | 7622 | hCoV-19/Luxembourg/LNS6321145/2020\|EPI_ISL_771133\| | ···················· | ···················· | ···················· | ···················· | ···················· | ···················· |
| AB | 7623 | hCoV-19/Luxembourg/LNS8161447/2020\|EPI_ISL_771134\| | ···················· | ···················· | ···················· | ···················· | ···················· | ···················· |
| AB | 7624 | hCoV-19/Luxembourg/LNS2992394/2020\|EPI_ISL_771135\| | ···················· | ···················· | ···················· | ···················· | ···················· | ···················· |
| AB | 7625 | hCoV-19/Luxembourg/LNS7201698/2020\|EPI_ISL_771136\| | ···················· | ···················· | ···················· | ···················· | ···················· | ···················· |
| AB | 7626 | hCoV-19/Luxembourg/LNS3633484/2020\|EPI_ISL_771137\| | ···················· | G··················· | ···················· | ···················· | ···················· | ···················· |

| | | | | | | | |
|---|---|---|---|---|---|---|---|
| AB | 7627 hCoV-19/Luxembourg/LNS2345895/2020\|EPI_ISL_771138\| | ···················· | ····A··············· | ···················· | ···················· | ···················· | ···················· |
| AB | 7628 hCoV-19/Luxembourg/LNS1072581/2020\|EPI_ISL_771139\| | ···················· | ····A··············· | ···················· | ···················· | ···················· | ···················· |
| AB | 7629 hCoV-19/Luxembourg/LNS8755341/2020\|EPI_ISL_771140\| | ···················· | ···················· | ···················· | ···················· | ···················· | ···················· |
| AB | 7630 hCoV-19/Luxembourg/LNS0171483/2020\|EPI_ISL_771141\| | ···················· | ···················· | ···················· | ···················· | ···················· | ···················· |
| AB | 7631 hCoV-19/Luxembourg/LNS2145340/2020\|EPI_ISL_771142\| | ···················· | ···················· | ···················· | ···················· | ···················· | ···················· |
| AB | 7632 hCoV-19/Luxembourg/LNS1180930/2020\|EPI_ISL_771143\| | ···················· | ···················· | ···················· | ···················· | ···················· | ···················· |
| AB | 7633 hCoV-19/Luxembourg/LNS5029028/2020\|EPI_ISL_771144\| | ···················· | ···················· | ···················· | ···················· | ···················· | ···················· |
| AB | 7634 hCoV-19/Luxembourg/LNS1761800/2020\|EPI_ISL_771145\| | ···················· | ···················· | ···················· | ···················· | ···················· | ···················· |
| AB | 7635 hCoV-19/Luxembourg/LNS6188449/2020\|EPI_ISL_771146\| | ···················· | ···················· | ···················· | ···················· | ···················· | ···················· |
| AB | 7636 hCoV-19/Luxembourg/LNS8256527/2020\|EPI_ISL_771147\| | ···················· | ···················· | ···················· | ···················· | ···················· | ···················· |
| AB | 7637 hCoV-19/Luxembourg/LNS5295339/2020\|EPI_ISL_771148\| | ···················· | ···················· | ···················· | ···················· | ···················· | ···················· |
| AB | 7638 hCoV-19/Luxembourg/LNS3342997/2020\|EPI_ISL_771149\| | ···················· | ···················· | ···················· | ···················· | ···················· | ···················· |
| AB | 7639 hCoV-19/Luxembourg/LNS9562893/2020\|EPI_ISL_771150\| | ···················· | ···················· | ···················· | ···················· | ···················· | ···················· |
| AB | 7640 hCoV-19/England/MILK-C90555/2020\|EPI_ISL_777200\|20 | ···················· | ···················· | ···················· | ···················· | ···················· | ···················· |
| AB | 7641 hCoV-19/England/CAMC-C8FEF4/2020\|EPI_ISL_777201\|20 | ···················· | ···················· | ···················· | ···················· | ···················· | ···················· |
| AB | 7642 hCoV-19/England/MILK-C90D02/2020\|EPI_ISL_777207\|20 | ···················· | ···················· | ···················· | ···················· | ···················· | ···················· |
| AB | 7643 hCoV-19/England/ALDP-C8F6B0/2020\|EPI_ISL_777211\|20 | ···················· | ···················· | ···················· | ···················· | ···················· | ···················· |
| AB | 7644 hCoV-19/England/ALDP-C8EA79/2020\|EPI_ISL_777213\|20 | ···················· | ···················· | ···················· | ···················· | ·····T·············· | ···················· |
| AB | 7645 hCoV-19/England/QEUH-C936B9/2020\|EPI_ISL_777215\|20 | ···················· | ···················· | ···················· | ···················· | ···················· | ···················· |
| AB | 7646 hCoV-19/England/ALDP-C8D0D2/2020\|EPI_ISL_777217\|20 | ···················· | ···················· | ···················· | ···················· | ···················· | ···················· |
| AB | 7647 hCoV-19/England/QEUH-C948D0/2020\|EPI_ISL_777261\|20 | ···················· | ···················· | ···················· | ···················· | ···················· | ···················· |
| AB | 7648 hCoV-19/England/ALDP-C8CC2A/2020\|EPI_ISL_777262\|20 | ···················· | ···················· | ···················· | ···················· | ···················· | ···················· |
| AB | 7649 hCoV-19/England/MILK-C9092C/2020\|EPI_ISL_777269\|20 | ···················· | ···················· | ···················· | ···················· | ···················· | ···················· |
| AB | 7650 hCoV-19/England/ALDP-C8EA3D/2020\|EPI_ISL_777270\|20 | ···················· | ···················· | ···················· | ···················· | ···················· | ···················· |
| AB | 7651 hCoV-19/England/CAMC-C91F38/2020\|EPI_ISL_777275\|20 | ···················· | ···················· | ···················· | ···················· | ···················· | ···················· |
| AB | 7652 hCoV-19/England/ALDP-C8C167/2020\|EPI_ISL_777276\|20 | ···················· | ···················· | ···················· | ···················· | ···················· | ···················· |
| AB | 7653 hCoV-19/England/ALDP-C8C64A/2020\|EPI_ISL_777278\|20 | ···················· | ···················· | ···················· | ···················· | ···················· | ···················· |
| AB | 7654 hCoV-19/England/QEUH-C91448/2020\|EPI_ISL_777284\|20 | ···················· | ·A·················· | ···················· | ···················· | ···················· | ··A················· |
| AB | 7655 hCoV-19/England/ALDP-C8CD09/2020\|EPI_ISL_777285\|20 | ···················· | ···················· | ···················· | ···················· | ···················· | ···················· |
| AB | 7656 hCoV-19/England/ALDP-C8F340/2020\|EPI_ISL_777286\|20 | ···················· | ·A·················· | ···················· | ···················· | ···················· | ···················· |
| AB | 7657 hCoV-19/England/QEUH-C93938/2020\|EPI_ISL_777290\|20 | ···················· | ···················· | ···················· | ···················· | ·····T·············· | ···················· |
| AB | 7658 hCoV-19/England/ALDP-C8D0F0/2020\|EPI_ISL_777291\|20 | ···················· | ···················· | ···················· | ···················· | ···················· | ···················· |
| AB | 7660 hCoV-19/England/CAMC-C92483/2020\|EPI_ISL_777293\|20 | ···················· | ···················· | ···················· | ···················· | ···················· | ···················· |
| AB | 7661 hCoV-19/England/MILK-C908B6/2020\|EPI_ISL_777295\|20 | ···················· | ···················· | ···················· | ···················· | ···················· | ···················· |
| AB | 7662 hCoV-19/England/ALDP-C8EA00/2020\|EPI_ISL_777296\|20 | ···················· | ···················· | ···················· | ···················· | ···················· | C··················· |
| AB | 7663 hCoV-19/England/ALDP-C8CC93/2020\|EPI_ISL_777297\|20 | ···················· | ···················· | ···················· | ···················· | ···················· | ···················· |
| AB | 7664 hCoV-19/England/ALDP-C8C404/2020\|EPI_ISL_777299\|20 | ···················· | ···················· | ···················· | ···················· | ···················· | ···················· |
| AB | 7665 hCoV-19/England/MILK-C6CBD8/2020\|EPI_ISL_778507\|20 | ···················· | ···················· | ···················· | ···················· | ···················· | ···················· |
| AB | 7666 hCoV-19/England/ALDP-C6B9DF/2020\|EPI_ISL_778510\|20 | ···················· | ···················· | ···················· | ···················· | ···················· | ···················· |
| AB | 7667 hCoV-19/England/ALDP-C6B975/2020\|EPI_ISL_778524\|20 | ···················· | ···················· | ···················· | ···················· | ···················· | ···················· |
| AB | 7668 hCoV-19/England/QEUH-C645D2/2020\|EPI_ISL_778527\|20 | ···················· | ···················· | ···················· | ···················· | ···················· | ···················· |
| AB | 7669 hCoV-19/England/ALDP-C6B9FD/2020\|EPI_ISL_778528\|20 | ···················· | ···················· | ···················· | ···················· | ···················· | ···················· |
| AB | 7670 hCoV-19/England/MILK-C6F6F2/2020\|EPI_ISL_778535\|20 | ···················· | ···················· | ···················· | ···················· | ···················· | ···················· |
| AB | 7671 hCoV-19/England/MILK-C6F64D/2020\|EPI_ISL_778536\|20 | ···················· | ···················· | ···················· | ···················· | ···················· | ···················· |
| AB | 7672 hCoV-19/England/MILK-C6CCA8/2020\|EPI_ISL_778537\|20 | ···················· | ···················· | ···················· | ···················· | ·····T·············· | ···················· |
| AB | 7673 hCoV-19/England/MILK-C6F461/2020\|EPI_ISL_778538\|20 | ···················· | ···················· | ···················· | ···················· | ···················· | ···················· |
| AB | 7674 hCoV-19/England/ALDP-C6BE67/2020\|EPI_ISL_778543\|20 | ···················· | ···················· | ···················· | ···················· | ···················· | ···················· |
| AB | 7675 hCoV-19/England/MILK-C6C9A1/2020\|EPI_ISL_778546\|20 | ···················· | ···················· | ···················· | ···················· | ···················· | ···················· |
| AB | 7676 hCoV-19/England/QEUH-C6444E/2020\|EPI_ISL_778548\|20 | ···················· | ···················· | ···················· | ···················· | ···················· | ···················· |
| AB | 7677 hCoV-19/England/ALDP-C6B1F5/2020\|EPI_ISL_778593\|20 | ···················· | ···················· | ···················· | ···················· | ···················· | ···················· |
| AB | 7678 hCoV-19/England/QEUH-C654F2/2020\|EPI_ISL_778594\|20 | ···················· | ···················· | ···················· | ···················· | ···················· | ···················· |
| AB | 7679 hCoV-19/England/MILK-C6F601/2020\|EPI_ISL_778595\|20 | ···················· | ···················· | ···················· | ···················· | ···················· | ···················· |
| AB | 7680 hCoV-19/England/ALDP-C6B720/2020\|EPI_ISL_778599\|20 | ···················· | ···················· | ···················· | ···················· | ···················· | ···················· |
| AB | 7681 hCoV-19/England/QEUH-CC5B55/2020\|EPI_ISL_760444\|20 | ···················· | ···················· | ···················· | ···················· | ···················· | ···················· |
| AB | 7682 hCoV-19/England/ALDP-CBDFEB/2020\|EPI_ISL_760445\|20 | ···················· | ···················· | ···················· | ···················· | ···················· | ···················· |
| AB | 7683 hCoV-19/England/QEUH-CC793B/2020\|EPI_ISL_760449\|20 | ···················· | ···················· | ···················· | ···················· | ·····T·············· | ···················· |
| AB | 7684 hCoV-19/England/QEUH-CC779B/2020\|EPI_ISL_760452\|20 | ···················· | ·A·················· | ···················· | ···················· | ···················· | ···················· |
| AB | 7685 hCoV-19/England/QEUH-CC5311/2020\|EPI_ISL_760453\|20 | ···················· | ···················· | ···················· | ···················· | ·····T·············· | ···················· |
| AB | 7686 hCoV-19/England/QEUH-CC7B9F/2020\|EPI_ISL_760454\|20 | ···················· | ···················· | ···················· | ···················· | ···················· | ···················· |
| AB | 7687 hCoV-19/England/QEUH-CC2639/2020\|EPI_ISL_760455\|20 | ···················· | ···················· | ···················· | ···················· | ···················· | ···················· |
| AB | 7688 hCoV-19/England/QEUH-CC5D40/2020\|EPI_ISL_760456\|20 | ···················· | ···················· | ···················· | ···················· | ···················· | ···················· |
| AB | 7689 hCoV-19/England/QEUH-CC7B53/2020\|EPI_ISL_760458\|20 | ···················· | ···················· | ···················· | ···················· | ·····T·············· | ···················· |
| AB | 7690 hCoV-19/England/QEUH-CC22EA/2020\|EPI_ISL_760462\|20 | ···················· | ···················· | ···················· | ···················· | ···················· | ···················· |
| AB | 7691 hCoV-19/England/QEUH-CC75DD/2020\|EPI_ISL_760464\|20 | ···················· | ···················· | ···················· | ···················· | ·····T·············· | ···················· |
| AB | 7692 hCoV-19/England/ALDP-CB55A5/2020\|EPI_ISL_761775\|20 | ···················· | ···················· | ···················· | ···················· | ···················· | ···················· |
| AB | 7693 hCoV-19/England/ALDP-CB511A/2020\|EPI_ISL_761776\|20 | ···················· | ···················· | ···················· | ···················· | ···················· | ···················· |
| AB | 7694 hCoV-19/England/ALDP-CB5253/2020\|EPI_ISL_761777\|20 | ···················· | ···················· | ···················· | ···················· | ···················· | ···················· |
| AB | 7695 hCoV-19/England/ALDP-CB5B58/2020\|EPI_ISL_761779\|20 | ···················· | ···················· | ···················· | ···················· | ···················· | ···················· |
| AB | 7696 hCoV-19/England/ALDP-CB4FE4/2020\|EPI_ISL_761780\|20 | ···················· | ···················· | ···················· | ···················· | ···················· | ···················· |
| AB | 7697 hCoV-19/England/ALDP-CB3783/2020\|EPI_ISL_761781\|20 | ···················· | ···················· | ···················· | ···················· | ···················· | ···················· |
| AB | 7698 hCoV-19/England/ALDP-CB65D1/2020\|EPI_ISL_761784\|20 | ···················· | ···················· | ···················· | ···················· | ···················· | ···················· |
| AB | 7699 hCoV-19/England/ALDP-CB6261/2020\|EPI_ISL_761786\|20 | ···················· | ···················· | ···················· | ···················· | ···················· | ···················· |
| AB | 7700 hCoV-19/England/ALDP-CB5BB2/2020\|EPI_ISL_761787\|20 | ···················· | ···················· | ···················· | ···················· | ···················· | ···················· |
| AB | 7701 hCoV-19/Wales/ALDP-CB39F6/2020\|EPI_ISL_761789\|2020 | ···················· | ···················· | ···················· | ···················· | ···················· | ···················· |
| AB | 7702 hCoV-19/England/ALDP-CB3CC0/2020\|EPI_ISL_761792\|20 | ···················· | ···················· | ···················· | ···················· | ···················· | ···················· |
| AB | 7703 hCoV-19/England/ALDP-CB65E0/2020\|EPI_ISL_761793\|20 | ···················· | ···················· | ···················· | ···················· | ···················· | ···················· |
| AB | 7706 hCoV-19/Wales/PHWC-4A675E/2020\|EPI_ISL_764065\|2020 | ···················· | ···················· | ···················· | ···················· | ···················· | ···················· |
| AB | 7707 hCoV-19/Sweden/20-33513/2020\|EPI_ISL_766600\|2020-1 | ···················· | ···················· | ···················· | ···················· | ···················· | ···················· |
| AB | 7708 hCoV-19/Sweden/20-33514/2020\|EPI_ISL_766601\|2020-1 | ···················· | ···················· | ···················· | ···················· | ···················· | ···················· |
| AB | 7709 hCoV-19/Sweden/20-33515/2020\|EPI_ISL_766602\|2020-1 | ···················· | ···················· | ···················· | ···················· | ···················· | ···················· |

AB 7710 hCoV-19/Sweden/20-33516/2020|EPI_ISL_766603|2020-1 .................... .................... .................... .................... .................... ....................
AB 7711 hCoV-19/Sweden/20-33518/2020|EPI_ISL_766604|2020-1 .................... .................... .................... .................... .................... ....................
AB 7712 hCoV-19/Sweden/20-33520/2020|EPI_ISL_766605|2020-1 .................... .................... .................... .................... .................... ....................
AB 7713 hCoV-19/Sweden/20-33523/2020|EPI_ISL_766606|2020-1 .................... .................... .................... .................... .................... ....................
AB 7714 hCoV-19/Sweden/20-33525/2020|EPI_ISL_766607|2020-1 .................... .................... .................... .................... .................... ....................
AB 7715 hCoV-19/Sweden/20-53767/2020|EPI_ISL_766608|2020-1 .................... .................... .................... .................... .................... ....................
AB 7716 hCoV-19/Sweden/20-53772/2020|EPI_ISL_766609|2020-1 .................... .................... .................... .................... .................... ....................
AB 7717 hCoV-19/Sweden/20-53776/2020|EPI_ISL_766610|2020-1 .................... .................... .................... .................... .................... ....................
AB 7718 hCoV-19/Sweden/20-53538/2020|EPI_ISL_766613|2020-1 .................... .................... .................... .................... .....T.............. ....................
AB 7719 hCoV-19/Sweden/20-53563/2020|EPI_ISL_766614|2020-1 .................... .................... .................... .................... .....T.............. ....................
AB 7720 hCoV-19/Sweden/20-53603/2020|EPI_ISL_766615|2020-1 .................... .................... .................... .................... .................... ....................
AB 7721 hCoV-19/Sweden/20-53606/2020|EPI_ISL_766616|2020-1 .................... .................... .................... .................... .................... ....................
AB 7722 hCoV-19/Sweden/20-53610/2020|EPI_ISL_766617|2020-1 .................... .................... .................... .................... .................... ....................
AB 7723 hCoV-19/Sweden/20-53612/2020|EPI_ISL_766618|2020-1 .................... .................... .................... .................... .................... ....................
AB 7724 hCoV-19/Sweden/20-53619/2020|EPI_ISL_766619|2020-1 .................... .................... .................... .................... .................... ....................
AB 7725 hCoV-19/Sweden/20-53620/2020|EPI_ISL_766620|2020-1 .................... .................... .................... .................... .................... ....................
AB 7726 hCoV-19/Sweden/20-53624/2020|EPI_ISL_766621|2020-1 .................... .................... .................... .................... .................... ....................
AB 7727 hCoV-19/Sweden/20-53636/2020|EPI_ISL_766622|2020-1 .................... .................... .................... .................... .................... ....................
AB 7728 hCoV-19/Sweden/20-53638/2020|EPI_ISL_766623|2020-1 .................... .................... .................... .................... .................... ....................
AB 7729 hCoV-19/Sweden/20-53640/2020|EPI_ISL_766624|2020-1 .................... .A.................. .................... .................... .................... ....................
AB 7730 hCoV-19/Sweden/20-52896/2020|EPI_ISL_766625|2020-0 .................... .................... .................... .................... .................... ....................
AB 7731 hCoV-19/Sweden/20-52897/2020|EPI_ISL_766626|2020-0 .................... .................... .................... .................... .................... ....................
AB 7732 hCoV-19/Sweden/20-52900/2020|EPI_ISL_766627|2020-0 .................... .................... .................... .................... .................... ....................
AB 7733 hCoV-19/Sweden/20-52903/2020|EPI_ISL_766628|2020-0 .................... .................... .................... .................... .................... ....................
AB 7734 hCoV-19/Sweden/20-52904/2020|EPI_ISL_766629|2020-0 .................... .................... .................... .................... .................... ....................
AB 7735 hCoV-19/Sweden/20-53719/2020|EPI_ISL_766630|2020-1 .................... .................... .................... .................... .................... ....................
AB 7736 hCoV-19/Sweden/20-53720/2020|EPI_ISL_766631|2020-1 .................... .................... .................... .................... .....T.............. ....................
AB 7737 hCoV-19/Sweden/20-53725/2020|EPI_ISL_766632|2020-1 .................... .................... .................... .................... .................... ....................
AB 7738 hCoV-19/Sweden/20-53761/2020|EPI_ISL_766633|2020-1 .................... .................... .................... .................... .................... ....................
AB 7739 hCoV-19/Sweden/20-53762/2020|EPI_ISL_766634|2020-1 .................... .................... .................... .................... .....T.............. ....................
AB 7740 hCoV-19/Sweden/20-53764/2020|EPI_ISL_766635|2020-1 .................... .................... .................... .................... .................... ....................
AB 7741 hCoV-19/Sweden/20-53766/2020|EPI_ISL_766636|2020-1 .................... .................... .................... .................... .....T.............. ....................
AB 7742 hCoV-19/Sweden/20-53770/2020|EPI_ISL_766637|2020-1 .................... .................... .................... .................... .................... ....................
AB 7743 hCoV-19/Sweden/20-53771/2020|EPI_ISL_766638|2020-1 .................... .................... .................... .................... .................... ....................
AB 7744 hCoV-19/Sweden/20-53773/2020|EPI_ISL_766639|2020-1 .................... .................... .................... .................... .................... ....................
AB 7745 hCoV-19/Sweden/20-53793/2020|EPI_ISL_766640|2020-1 .................... .................... .................... .................... .................... ....................
AB 7746 hCoV-19/Sweden/20-53801/2020|EPI_ISL_766641|2020-1 .................... .................... .................... .................... .................... ....................
AB 7747 hCoV-19/Sweden/20-24578/2020|EPI_ISL_766642|2020-1 .................... .................... .................... .................... .................... ....................
AB 7748 hCoV-19/Sweden/20-24736/2020|EPI_ISL_766643|2020-1 .................... .................... .................... .................... .................... ....................
AB 7749 hCoV-19/Sweden/20-33272/2020|EPI_ISL_766644|2020-1 .................... .................... .................... .................... .................... ....................
AB 7750 hCoV-19/Sweden/20-53575/2020|EPI_ISL_766645|2020-1 .................... .........A.......... .................... .................... .................... ....................
AB 7751 hCoV-19/USA/TX-DSHS-2028/2020|EPI_ISL_766687|2020- .................... .................... .................... .................... .................... ....................
AB 7752 hCoV-19/USA/TX-DSHS-2049/2020|EPI_ISL_766688|2020- .................... .................... .................... .................... .................... ....................
AB 7753 hCoV-19/USA/TX-DSHS-2050/2020|EPI_ISL_766689|2020- .................... .................... .................... .................... .................... ....................
AB 7754 hCoV-19/USA/TX-DSHS-2056/2020|EPI_ISL_766690|2020- .................... .................... .................... .................... .................... ....................
AB 7755 hCoV-19/Australia/NSW1334/2020|EPI_ISL_767900|2020 .................... .................... .................... .................... .................... ....................
AB 7756 hCoV-19/Australia/NSW1335/2020|EPI_ISL_767901|2020 .................... .................... .................... .................... .................... ....................
AB 7757 hCoV-19/Australia/NSW1336/2020|EPI_ISL_767902|2020 .................... .................... .................... .................... .................... ....................
AB 7758 hCoV-19/Australia/NSW1337/2020|EPI_ISL_767903|2020 .................... .................... .................... .................... .................... ....................
AB 7759 hCoV-19/Australia/NSW1338/2020|EPI_ISL_767904|2020 .................... .................... .................... .................... .................... ....................
AB 7760 hCoV-19/Australia/NSW1339/2020|EPI_ISL_767905|2020 .................... .................... .................... .................... .................... ....................
AB 7761 hCoV-19/Australia/NSW1340/2020|EPI_ISL_767906|2020 .................... .................... .................... .................... .................... ....................
AB 7762 hCoV-19/Australia/NSW1341/2020|EPI_ISL_767907|2020 .................... .................... .................... .................... .................... ....................
AB 7763 hCoV-19/Australia/NSW1342/2020|EPI_ISL_767908|2020 .................... .................... .................... .................... .................... ....................
AB 7764 hCoV-19/Australia/NSW1343/2020|EPI_ISL_767909|2020 .................... .................... .................... .................... .................... ....................
AB 7765 hCoV-19/Australia/NSW1344/2020|EPI_ISL_767910|2020 .................... .................... .................... .................... .................... ....................
AB 7766 hCoV-19/Australia/NSW1345/2020|EPI_ISL_767911|2020 .................... .................... .................... .................... .................... ....................
AB 7767 hCoV-19/Australia/NSW1346/2020|EPI_ISL_767912|2020 .................... .................... .................... .................... .................... ....................
AB 7768 hCoV-19/Australia/NSW1347/2020|EPI_ISL_767913|2020 .................... .................... .................... .................... .................... ....................
AB 7769 hCoV-19/Australia/NSW1348/2020|EPI_ISL_767914|2020 .................... .................... .................... .................... .................... ....................
AB 7770 hCoV-19/Australia/NSW1349/2020|EPI_ISL_767915|2020 .................... .................... .................... .................... .................... ....................
AB 7771 hCoV-19/Australia/NSW1350/2020|EPI_ISL_767916|2020 .................... .................... .................... .................... .................... ....................
AB 7772 hCoV-19/Australia/NSW1351/2020|EPI_ISL_767917|2020 .................... .................... .................... .................... .................... ....................
AB 7773 hCoV-19/Australia/NSW1352/2020|EPI_ISL_767918|2020 .................... .................... .................... .................... .................... ....................
AB 7774 hCoV-19/Australia/NSW1353/2020|EPI_ISL_767919|2020 .................... .................... .................... .................... .................... ....................
AB 7775 hCoV-19/Australia/NSW1354/2020|EPI_ISL_767920|2020 .................... .................... .................... .................... .................... ....................
AB 7776 hCoV-19/Australia/NSW1355/2020|EPI_ISL_767921|2020 .................... .................... .................... .................... .................... ....................
AB 7777 hCoV-19/Australia/NSW1356/2020|EPI_ISL_767922|2020 .................... .................... .................... .................... .................... ....................
AB 7778 hCoV-19/Australia/NSW1357/2020|EPI_ISL_767923|2020 .................... .................... .................... .................... .................... ....................
AB 7779 hCoV-19/Australia/NSW1358/2020|EPI_ISL_767924|2020 .................... .................... .................... .................... .................... ....................
AB 7780 hCoV-19/Australia/NSW1359/2020|EPI_ISL_767925|2020 .................... .................... .................... .................... .................... ....................
AB 7781 hCoV-19/Australia/NSW1360/2020|EPI_ISL_767926|2020 .................... .................... .................... .................... .................... ....................
AB 7782 hCoV-19/Australia/NSW1361/2020|EPI_ISL_767927|2020 .................... .................... .................... .................... .................... ....................
AB 7783 hCoV-19/Australia/NSW1362/2020|EPI_ISL_767928|2020 .................... .................... .................... .................... .................... ....................
AB 7784 hCoV-19/Australia/NSW1363/2020|EPI_ISL_767929|2020 .................... .................... .................... .................... .................... ....................
AB 7785 hCoV-19/Australia/NSW1364/2020|EPI_ISL_767930|2020 .................... .................... .................... .................... .................... ....................
AB 7786 hCoV-19/Australia/NSW1365/2020|EPI_ISL_767931|2020 .................... .................... .................... .................... .................... ....................
AB 7787 hCoV-19/Australia/NSW1366/2020|EPI_ISL_767932|2020 .................... .................... .................... .................... .................... ....................
AB 7788 hCoV-19/Australia/NSW1367/2020|EPI_ISL_767933|2020 .................... .................... .................... .................... .................... ....................
AB 7789 hCoV-19/Australia/NSW1368/2020|EPI_ISL_767934|2020 .................... .................... .................... .................... .................... ....................

| | | | | | | | |
|---|---|---|---|---|---|---|---|
| AB | 7790 hCoV-19/Australia/NSW1369/2020\|EPI_ISL_767935\|2020 | ···················· | ···················· | ····················· | ···················· | ···················· | ···················· |
| AB | 7791 hCoV-19/Australia/NSW1370/2020\|EPI_ISL_767936\|2020 | ···················· | ···················· | ····················· | ···················· | ···················· | ···················· |
| AB | 7792 hCoV-19/Australia/NSW1371/2021\|EPI_ISL_767937\|2021 | ···················· | ···················· | ····················· | ···················· | ···················· | ···················· |
| AB | 7793 hCoV-19/Australia/NSW1372/2021\|EPI_ISL_767938\|2021 | ···················· | ···················· | ····················· | ···················· | ···················· | ···················· |
| AB | 7794 hCoV-19/Australia/NSW1373/2021\|EPI_ISL_767939\|2021 | ···················· | ···················· | ····················· | ···················· | ···················· | ···················· |
| AB | 7795 hCoV-19/Australia/NSW1374/2021\|EPI_ISL_767940\|2021 | ···················· | ···················· | ····················· | ···················· | ···················· | ···················· |
| AB | 7796 hCoV-19/Switzerland/AG-ETHZ-420517/2020\|EPI_ISL_76 | ···················· | ···················· | ····················· | ···················· | ···················· | ···················· |
| AB | 7800 hCoV-19/Switzerland/AG-ETHZ-420113/2020\|EPI_ISL_76 | ···················· | ···················· | ····················· | ···················· | ···················· | ···················· |
| AB | 7802 hCoV-19/Luxembourg/LNS2162254/2020\|EPI_ISL_771100\| | ···················· | ···················· | ····················· | ···················· | ···················· | ···················· |
| AB | 7803 hCoV-19/Luxembourg/LNS6722121/2020\|EPI_ISL_771101\| | ···················· | ···················· | ····················· | ···················· | ······T············· | ···················· |
| AB | 7804 hCoV-19/Luxembourg/LNS6205753/2020\|EPI_ISL_771102\| | ···················· | ···················· | ····················· | ···················· | ···················· | ···················· |
| AB | 7805 hCoV-19/Luxembourg/LNS6982714/2020\|EPI_ISL_771106\| | ···················· | ···················· | ····················· | ···················· | ···················· | ···················· |
| AB | 7806 hCoV-19/Luxembourg/LNS2437179/2020\|EPI_ISL_771107\| | ···················· | ···················· | ····················· | ···················· | ···················· | ···················· |
| AB | 7807 hCoV-19/Luxembourg/LNS6774334/2020\|EPI_ISL_771108\| | ···················· | G··················· | ····················· | ···················· | ···················· | ···················· |
| AB | 7808 hCoV-19/Luxembourg/LNS7400495/2020\|EPI_ISL_771109\| | ···················· | G··················· | ····················· | ···················· | ···················· | ···················· |
| AB | 7809 hCoV-19/Luxembourg/LNS8042017/2020\|EPI_ISL_771110\| | ···················· | ···················· | ····················· | ···················· | ···················· | ···················· |
| AB | 7810 hCoV-19/Luxembourg/LNS2492377/2020\|EPI_ISL_771111\| | ···················· | ···················· | ····················· | ···················· | ···················· | ···················· |
| AB | 7811 hCoV-19/Luxembourg/LNS2211019/2020\|EPI_ISL_771112\| | ···················· | ···················· | ····················· | ···················· | ···················· | ···················· |
| AB | 7812 hCoV-19/USA/CO-CDPHE-2100013054/2020\|EPI_ISL_77118 | ···················· | ···················· | ····················· | ···················· | ···················· | ···················· |
| AB | 7813 hCoV-19/USA/CO-CDPHE-2008022512/2020\|EPI_ISL_77119 | ···················· | ···················· | ····················· | ···················· | ···················· | ···················· |
| AB | 7814 hCoV-19/USA/CO-CDPHE-2100028126/2020\|EPI_ISL_77119 | ···················· | ···················· | ····················· | ···················· | ···················· | ···················· |
| AB | 7815 hCoV-19/Germany/HH-hpi-p1492/2020\|EPI_ISL_776000\|2 | ···················· | ···················· | ····················· | ···················· | ···················· | ···················· |
| AB | 7816 hCoV-19/Germany/HH-hpi-p1493/2020\|EPI_ISL_776001\|2 | ···················· | ···················· | ····················· | ···················· | ···················· | ···················· |
| AB | 7817 hCoV-19/Germany/HH-hpi-p1494/2020\|EPI_ISL_776002\|2 | ···················· | ···················· | ····················· | ···················· | ···················· | ···················· |
| AB | 7818 hCoV-19/Germany/HH-hpi-p1497/2020\|EPI_ISL_776003\|2 | ···················· | ···················· | ····················· | ···················· | ···················· | ···················· |
| AB | 7819 hCoV-19/Germany/HH-hpi-p1498/2020\|EPI_ISL_776004\|2 | ···················· | ·············A······ | ····················· | ···················· | ···················· | ···················· |
| AB | 7820 hCoV-19/Germany/HH-hpi-p1499/2020\|EPI_ISL_776005\|2 | ···················· | ···················· | ····················· | ···················· | ···················· | ···················· |
| AB | 7821 hCoV-19/Germany/HH-hpi-p1500/2020\|EPI_ISL_776006\|2 | ···················· | ···················· | ····················· | ···················· | ···················· | ···················· |
| AB | 7822 hCoV-19/Germany/HH-hpi-p1501/2020\|EPI_ISL_776007\|2 | ···················· | ···················· | ····················· | ···················· | ···················· | ···················· |
| AB | 7823 hCoV-19/Germany/HH-hpi-p1502/2020\|EPI_ISL_776008\|2 | ···················· | ···················· | ····················· | ···················· | ···················· | ···················· |
| AB | 7824 hCoV-19/Germany/HH-hpi-p1503/2020\|EPI_ISL_776009\|2 | ···················· | ···················· | ····················· | ···················· | ···················· | ···················· |
| AB | 7825 hCoV-19/Germany/HH-hpi-p1504/2020\|EPI_ISL_776010\|2 | ···················· | ···················· | ····················· | ···················· | ···················· | ···················· |
| AB | 7826 hCoV-19/Germany/HH-hpi-p1506/2020\|EPI_ISL_776011\|2 | ···················· | ···················· | ····················· | ···················· | ···················· | ···················· |
| AB | 7827 hCoV-19/Germany/HH-hpi-p1507/2020\|EPI_ISL_776012\|2 | ···················· | ···················· | ····················· | ···················· | ···················· | ···················· |
| AB | 7828 hCoV-19/Germany/HH-hpi-p1508/2020\|EPI_ISL_776013\|2 | ···················· | ···················· | ····················· | ···················· | ···················· | ···················· |
| AB | 7829 hCoV-19/Germany/HH-hpi-p1509/2020\|EPI_ISL_776014\|2 | ···················· | ···················· | ····················· | ···················· | ···················· | ···················· |
| AB | 7830 hCoV-19/Germany/HH-hpi-p1510/2020\|EPI_ISL_776015\|2 | ···················· | ···················· | ····················· | ···················· | ···················· | ···················· |
| AB | 7832 hCoV-19/Germany/HH-hpi-p1514/2020\|EPI_ISL_776017\|2 | ···················· | ···················· | ····················· | ···················· | ···················· | ···················· |
| AB | 7833 hCoV-19/Germany/HH-hpi-p1516/2020\|EPI_ISL_776018\|2 | ···················· | ···················· | ····················· | ···················· | ···················· | ···················· |
| AB | 7834 hCoV-19/Germany/HH-hpi-p1517/2020\|EPI_ISL_776019\|2 | ···················· | ·············A······ | ····················· | ···················· | ···················· | ···················· |
| AB | 7835 hCoV-19/Germany/HH-hpi-p1519/2020\|EPI_ISL_776020\|2 | ···················· | ···················· | ····················· | ···················· | ···················· | ···················· |
| AB | 7836 hCoV-19/Germany/HH-hpi-p1520/2020\|EPI_ISL_776021\|2 | ···················· | ···················· | ····················· | ···················· | ···················· | ···················· |
| AB | 7837 hCoV-19/Germany/HH-hpi-p1521/2020\|EPI_ISL_776022\|2 | ···················· | ···················· | ····················· | ···················· | ···················· | ···················· |
| AB | 7838 hCoV-19/Germany/HH-hpi-p1522/2020\|EPI_ISL_776023\|2 | ···················· | ···················· | ····················· | ···················· | ···················· | ···················· |
| AB | 7839 hCoV-19/Germany/HH-hpi-p1524/2020\|EPI_ISL_776024\|2 | ···················· | ···················· | ····················· | ···················· | ···················· | ···················· |
| AB | 7840 hCoV-19/Germany/HH-hpi-p1525/2020\|EPI_ISL_776025\|2 | ···················· | ···················· | ····················· | ···················· | ···················· | ···················· |
| AB | 7841 hCoV-19/Germany/HH-hpi-p1527/2020\|EPI_ISL_776026\|2 | ···················· | ···················· | ····················· | ···················· | ···················· | ···················· |
| AB | 7842 hCoV-19/Germany/HH-hpi-p1529/2020\|EPI_ISL_776027\|2 | ···················· | ···················· | ····················· | ···················· | ···················· | ···················· |
| AB | 7843 hCoV-19/Germany/HH-hpi-p1533/2020\|EPI_ISL_776028\|2 | ···················· | ···················· | ····················· | ···················· | ···················· | ···················· |
| AB | 7844 hCoV-19/Germany/HH-hpi-p1534/2020\|EPI_ISL_776029\|2 | ···················· | ···················· | ····················· | ···················· | ···················· | ···················· |
| AB | 7845 hCoV-19/Germany/HH-hpi-p1535/2020\|EPI_ISL_776030\|2 | ···················· | ···················· | ····················· | ···················· | ···················· | ···················· |
| AB | 7847 hCoV-19/Germany/HH-hpi-p1538/2020\|EPI_ISL_776032\|2 | ···················· | ···················· | ····················· | ···················· | ···················· | ···················· |
| AB | 7848 hCoV-19/Germany/HH-hpi-p1541/2020\|EPI_ISL_776033\|2 | ···················· | ···················· | ····················· | ···················· | ···················· | ···················· |
| AB | 7849 hCoV-19/Germany/HH-hpi-p1543/2020\|EPI_ISL_776034\|2 | ···················· | ···················· | ····················· | ···················· | ···················· | ···················· |
| AB | 7850 hCoV-19/Germany/HH-hpi-p1544/2020\|EPI_ISL_776035\|2 | ···················· | ···················· | ····················· | ···················· | ···················· | ···················· |
| AB | 7851 hCoV-19/Germany/HH-hpi-p1550/2020\|EPI_ISL_776036\|2 | ···················· | ···················· | ····················· | ···················· | ···················· | ···················· |
| AB | 7852 hCoV-19/Germany/HH-hpi-p1551/2020\|EPI_ISL_776037\|2 | ···················· | ···················· | ····················· | ···················· | ···················· | ···················· |
| AB | 7853 hCoV-19/Germany/HH-hpi-p1554/2020\|EPI_ISL_776038\|2 | ···················· | ···················· | ····················· | ···················· | ···················· | ···················· |
| AB | 7854 hCoV-19/Germany/HH-hpi-p1555/2020\|EPI_ISL_776039\|2 | ···················· | ···················· | ····················· | ···················· | ···················· | ···················· |
| AB | 7855 hCoV-19/Germany/HH-hpi-p1558/2020\|EPI_ISL_776040\|2 | ···················· | ···················· | ····················· | ···················· | ···················· | ···················· |
| AB | 7856 hCoV-19/Germany/HH-hpi-p1559/2020\|EPI_ISL_776041\|2 | ···················· | ···················· | ····················· | ···················· | ···················· | ···················· |
| AB | 7857 hCoV-19/Germany/HH-hpi-p1560/2020\|EPI_ISL_776042\|2 | ···················· | ···················· | ····················· | ···················· | ···················· | ···················· |
| AB | 7858 hCoV-19/Germany/HH-hpi-p1561/2020\|EPI_ISL_776043\|2 | ···················· | ···················· | ····················· | ···················· | ···················· | ···················· |
| AB | 7859 hCoV-19/Germany/HH-hpi-p1563/2020\|EPI_ISL_776044\|2 | ···················· | ···················· | ····················· | ···················· | ···················· | ···················· |
| AB | 7860 hCoV-19/Germany/HH-hpi-p1565/2020\|EPI_ISL_776045\|2 | ···················· | ···················· | ····················· | ···················· | ···················· | ···················· |
| AB | 7861 hCoV-19/Germany/HH-hpi-p1567/2020\|EPI_ISL_776046\|2 | ···················· | ···················· | ····················· | ···················· | ···················· | ···················· |
| AB | 7862 hCoV-19/Germany/HH-hpi-p1568/2020\|EPI_ISL_776047\|2 | ···················· | ···················· | ····················· | ···················· | ···················· | ···················· |
| AB | 7863 hCoV-19/Germany/HH-hpi-p1569/2020\|EPI_ISL_776048\|2 | ···················· | ···················· | ····················· | ···················· | ···················· | ···················· |
| AB | 7864 hCoV-19/Germany/HH-hpi-p1570/2020\|EPI_ISL_776049\|2 | ···················· | ···················· | ····················· | ···················· | ···················· | ···················· |
| AB | 7865 hCoV-19/England/QEUH-C9368C/2020\|EPI_ISL_777302\|20 | ···················· | ···················· | ····················· | ···················· | ···················· | ···················· |
| AB | 7866 hCoV-19/England/MILK-C90B9F/2020\|EPI_ISL_777304\|20 | ···················· | ···················· | ····················· | ···················· | ···················· | ···················· |
| AB | 7867 hCoV-19/England/QEUH-C93A44/2020\|EPI_ISL_777305\|20 | ···················· | ···················· | ····················· | ···················· | ···················· | ···················· |
| AB | 7868 hCoV-19/England/QEUH-C948B2/2020\|EPI_ISL_777306\|20 | ···················· | ···················· | ····················· | ···················· | ···················· | ···················· |
| AB | 7869 hCoV-19/England/ALDP-C8D0C3/2020\|EPI_ISL_777308\|20 | ···················· | ···················· | ····················· | ···················· | ···················· | ···················· |
| AB | 7870 hCoV-19/England/MILK-C906E9/2020\|EPI_ISL_777313\|20 | ···················· | ···················· | ····················· | ···················· | ···················· | ···················· |
| AB | 7871 hCoV-19/England/ALDP-C8FD8E/2020\|EPI_ISL_777314\|20 | ···················· | ···················· | ····················· | ···················· | ···················· | ···················· |
| AB | 7872 hCoV-19/England/QEUH-C93640/2020\|EPI_ISL_777316\|20 | ···················· | ···················· | ····················· | ···················· | ···················· | ···················· |
| AB | 7873 hCoV-19/England/ALDP-C8BCB2/2020\|EPI_ISL_777319\|20 | ···················· | ···················· | ····················· | ···················· | ···················· | ···················· |
| AB | 7874 hCoV-19/England/CAMC-C98D28/2020\|EPI_ISL_777332\|20 | ···················· | ···················· | ····················· | ···················· | ···················· | ···················· |
| AB | 7875 hCoV-19/England/CAMC-C9863C/2020\|EPI_ISL_777343\|20 | ···················· | ···················· | ····················· | ···················· | ···················· | ···················· |

| | | | | | | | | |
|---|---|---|---|---|---|---|---|---|
| AB | 7876 | hCoV-19/England/CAMC-C9898E/2020\|EPI_ISL_777351\|20 | .................... | .................... | ..................... | .................... | .................... | .................... |
| AB | 7877 | hCoV-19/England/CAMC-C98AF4/2020\|EPI_ISL_777359\|20 | .................... | .................... | ..................... | .................... | .................... | .................... |
| AB | 7878 | hCoV-19/England/CAMC-C98854/2020\|EPI_ISL_777360\|20 | .................... | .................... | ..................... | .................... | .................... | .................... |
| AB | 7879 | hCoV-19/England/QEUH-C93ACC/2020\|EPI_ISL_777363\|20 | .................... | .................... | ..................... | .................... | .................... | .................... |
| AB | 7880 | hCoV-19/England/CAMC-C9939E/2020\|EPI_ISL_777366\|20 | .................... | .................... | ..................... | .................... | .................... | .................... |
| AB | 7881 | hCoV-19/England/CAMC-C993AD/2020\|EPI_ISL_777367\|20 | .................... | .................... | ..................... | .................... | .................... | .................... |
| AB | 7882 | hCoV-19/England/QEUH-C93FBE/2020\|EPI_ISL_777373\|20 | .................... | .................... | ..................... | .................... | .................... | .................... |
| AB | 7883 | hCoV-19/England/ALDP-C8D9E6/2020\|EPI_ISL_777375\|20 | .................... | .................... | ..................... | .................... | ......T............. | .................... |
| AB | 7884 | hCoV-19/England/QEUH-C93F81/2020\|EPI_ISL_777376\|20 | .................... | .................... | ..................... | .................... | .................... | .................... |
| AB | 7885 | hCoV-19/England/MILK-C6CFBE/2020\|EPI_ISL_778600\|20 | .................... | .A.................. | ..................... | .................... | .................... | .................... |
| AB | 7886 | hCoV-19/England/MILK-C6F698/2020\|EPI_ISL_778602\|20 | .................... | .................... | ..................... | .................... | .................... | .................... |
| AB | 7887 | hCoV-19/England/MILK-C6F689/2020\|EPI_ISL_778603\|20 | .................... | .................... | ..................... | .................... | .................... | .................... |
| AB | 7888 | hCoV-19/England/MILK-C6F63E/2020\|EPI_ISL_778611\|20 | .................... | .................... | ..................... | .................... | .................... | .................... |
| AB | 7889 | hCoV-19/England/ALDP-C6BA09/2020\|EPI_ISL_778614\|20 | .................... | .................... | ..................... | .................... | .................... | .................... |
| AB | 7890 | hCoV-19/England/ALDP-C6B8B4/2020\|EPI_ISL_778615\|20 | .................... | .................... | ..................... | .................... | .................... | .................... |
| AB | 7891 | hCoV-19/England/ALDP-C6B8D2/2020\|EPI_ISL_778630\|20 | .................... | .................... | ..................... | .................... | .................... | .................... |
| AB | 7892 | hCoV-19/England/ALDP-C6A18D/2020\|EPI_ISL_778635\|20 | .................... | .................... | ..................... | .................... | .................... | .................... |
| AB | 7893 | hCoV-19/England/MILK-C6F73B/2020\|EPI_ISL_778636\|20 | .................... | .................... | ..................... | .................... | .................... | .................... |
| AB | 7894 | hCoV-19/England/MILK-C6F838/2020\|EPI_ISL_778637\|20 | .................... | .................... | ..................... | .................... | .................... | .................... |
| AB | 7895 | hCoV-19/England/MILK-C60D74/2020\|EPI_ISL_778641\|20 | .................... | .................... | ..................... | .................... | .................... | .................... |
| AB | 7896 | hCoV-19/England/MILK-C51927/2020\|EPI_ISL_778643\|20 | .................... | .................... | ..................... | .................... | .................... | .................... |
| AB | 7897 | hCoV-19/Spain/M-20115914/2020\|EPI_ISL_778644\|2020- | .................... | .................... | ..................... | .................... | .................... | .................... |
| AB | 7898 | hCoV-19/Spain/M-20105262/2020\|EPI_ISL_778645\|2020- | .................... | .................... | ..................... | .................... | .................... | .................... |
| AB | 7899 | hCoV-19/Italy/CAM-TIGEM-589/2020\|EPI_ISL_778646\|20 | .................... | .................... | ..................... | .................... | .................... | .................... |
| AB | 7900 | hCoV-19/Italy/CAM-TIGEM-636/2020\|EPI_ISL_778647\|20 | .................... | .................... | ..................... | .................... | .................... | .................... |
| AB | 7901 | hCoV-19/Italy/CAM-TIGEM-600/2020\|EPI_ISL_778648\|20 | .................... | .................... | ..................... | .................... | .................... | .................... |
| AB | 7902 | hCoV-19/Italy/CAM-TIGEM-601/2020\|EPI_ISL_778649\|20 | .................... | .................... | ..................... | .................... | .................... | .................... |
| AB | 7903 | hCoV-19/Italy/CAM-TIGEM-581/2020\|EPI_ISL_778659\|20 | .................... | .................... | ..................... | .................... | .................... | .................... |
| AB | 7904 | hCoV-19/Italy/CAM-TIGEM-582/2020\|EPI_ISL_778660\|20 | .................... | .................... | ..................... | .................... | .................... | .................... |
| AB | 7905 | hCoV-19/Italy/CAM-TIGEM-583/2020\|EPI_ISL_778661\|20 | .................... | .................... | ..................... | .................... | .................... | .................... |
| AB | 7906 | hCoV-19/Italy/CAM-TIGEM-595/2020\|EPI_ISL_778662\|20 | .................... | .................... | ..................... | .................... | .................... | .................... |
| AB | 7907 | hCoV-19/Italy/CAM-TIGEM-590/2020\|EPI_ISL_778663\|20 | .................... | .................... | ..................... | .................... | .................... | .................... |
| AB | 7908 | hCoV-19/Italy/CAM-TIGEM-591/2020\|EPI_ISL_778664\|20 | .................... | .................... | ..................... | .................... | .................... | .................... |
| AB | 7909 | hCoV-19/Italy/CAM-TIGEM-592/2020\|EPI_ISL_778665\|20 | .................... | .................... | ..................... | .................... | .................... | .................... |
| AB | 7910 | hCoV-19/Italy/CAM-TIGEM-588/2020\|EPI_ISL_778666\|20 | .................... | .................... | ..................... | .................... | .................... | .................... |
| AB | 7911 | hCoV-19/Italy/CAM-TIGEM-598/2020\|EPI_ISL_778667\|20 | .................... | .................... | ..................... | .................... | .................... | .................... |
| AB | 7912 | hCoV-19/Italy/CAM-TIGEM-599/2020\|EPI_ISL_778669\|20 | .................... | .................... | ..................... | .................... | .................... | .................... |
| AB | 7913 | hCoV-19/Italy/CAM-TIGEM-606/2020\|EPI_ISL_778670\|20 | .................... | .................... | ..................... | .................... | .................... | .................... |
| AB | 7914 | hCoV-19/Italy/CAM-TIGEM-608/2020\|EPI_ISL_778671\|20 | .................... | .................... | ..................... | .................... | .................... | .................... |
| AB | 7915 | hCoV-19/Italy/CAM-TIGEM-609/2020\|EPI_ISL_778673\|20 | .................... | .................... | ..................... | .................... | .................... | .................... |
| AB | 7916 | hCoV-19/Italy/CAM-TIGEM-610/2020\|EPI_ISL_778674\|20 | .................... | .................... | ..................... | .................... | .................... | .................... |
| AB | 7917 | hCoV-19/Italy/CAM-TIGEM-618/2020\|EPI_ISL_778676\|20 | .................... | .................... | ..................... | .................... | .................... | .................... |
| AB | 7918 | hCoV-19/Italy/CAM-TIGEM-619/2020\|EPI_ISL_778677\|20 | .................... | .................... | ..................... | .................... | .................... | .................... |
| AB | 7919 | hCoV-19/Italy/CAM-TIGEM-620/2020\|EPI_ISL_778678\|20 | .................... | .................... | ..................... | .................... | .................... | .................... |
| AB | 7920 | hCoV-19/Italy/CAM-TIGEM-621/2020\|EPI_ISL_778679\|20 | .................... | .................... | ..................... | .................... | .................... | .................... |
| AB | 7921 | hCoV-19/Italy/CAM-TIGEM-623/2020\|EPI_ISL_778680\|20 | .................... | .................... | ..................... | .................... | .................... | .................... |
| AB | 7922 | hCoV-19/Italy/CAM-TIGEM-616/2020\|EPI_ISL_778681\|20 | .................... | .................... | ..................... | .................... | .................... | .................... |
| AB | 7923 | hCoV-19/Italy/CAM-TIGEM-628/2020\|EPI_ISL_778682\|20 | .................... | .................... | ..................... | .................... | .................... | .................... |
| AB | 7924 | hCoV-19/Italy/CAM-TIGEM-630/2020\|EPI_ISL_778683\|20 | .................... | .................... | ..................... | .................... | .................... | .................... |
| AB | 7925 | hCoV-19/Italy/CAM-TIGEM-631/2020\|EPI_ISL_778684\|20 | .................... | .................... | ..................... | .................... | .................... | .................... |
| AB | 7926 | hCoV-19/Italy/CAM-TIGEM-629/2020\|EPI_ISL_778685\|20 | .................... | .................... | ..................... | .................... | .................... | .................... |
| AB | 7927 | hCoV-19/Italy/CAM-TIGEM-665/2020\|EPI_ISL_778686\|20 | .................... | .................... | ..................... | .................... | .................... | .................... |
| AB | 7928 | hCoV-19/Italy/CAM-TIGEM-666/2020\|EPI_ISL_778687\|20 | .................... | .................... | ..................... | .................... | .................... | .................... |
| AB | 7929 | hCoV-19/Italy/CAM-TIGEM-668/2020\|EPI_ISL_778689\|20 | .................... | .................... | ..................... | .................... | .................... | .................... |
| AB | 7930 | hCoV-19/Italy/CAM-TIGEM-669/2020\|EPI_ISL_778690\|20 | .................... | .................... | ..................... | .................... | .................... | .................... |
| AB | 7931 | hCoV-19/Italy/CAM-TIGEM-417/2020\|EPI_ISL_778691\|20 | .................... | .................... | ..................... | .................... | .................... | .................... |
| AB | 7932 | hCoV-19/Italy/CAM-TIGEM-397/2020\|EPI_ISL_778692\|20 | .................... | .................... | ..................... | .................... | ......T............. | .................... |
| AB | 7933 | hCoV-19/Italy/CAM-TIGEM-389/2020\|EPI_ISL_778693\|20 | .................... | .................... | ..................... | .................... | ......T............. | .................... |
| AB | 7934 | hCoV-19/Italy/CAM-TIGEM-390/2020\|EPI_ISL_778695\|20 | .................... | .................... | ..................... | .................... | .................... | .................... |
| AB | 7935 | hCoV-19/Italy/CAM-TIGEM-564/2020\|EPI_ISL_778696\|20 | .................... | .................... | ..................... | .................... | .................... | .................... |
| AB | 7936 | hCoV-19/Italy/CAM-TIGEM-568/2020\|EPI_ISL_778699\|20 | .................... | .................... | ..................... | .................... | .................... | .................... |
| AB | 7937 | hCoV-19/England/ALDP-CC005B/2020\|EPI_ISL_760500\|20 | .................... | .................... | ..................... | .................... | .................... | .................... |
| AB | 7938 | hCoV-19/England/QEUH-CC5B82/2020\|EPI_ISL_760502\|20 | .................... | .................... | ..................... | .................... | .................... | .................... |
| AB | 7939 | hCoV-19/England/QEUH-CC5B64/2020\|EPI_ISL_760503\|20 | .................... | .................... | ..................... | .................... | .................... | .................... |
| AB | 7940 | hCoV-19/England/QEUH-CC2350/2020\|EPI_ISL_760505\|20 | .................... | .................... | ..................... | .................... | .................... | .................... |
| AB | 7941 | hCoV-19/England/QEUH-CC7625/2020\|EPI_ISL_760506\|20 | .................... | .................... | ..................... | .................... | .................... | .................... |
| AB | 7942 | hCoV-19/England/ALDP-CC08F9/2020\|EPI_ISL_760509\|20 | .................... | .................... | ..................... | .................... | ......T............. | .................... |
| AB | 7943 | hCoV-19/England/QEUH-CC5DB9/2020\|EPI_ISL_760513\|20 | .................... | .................... | ..................... | .................... | .................... | .................... |
| AB | 7944 | hCoV-19/England/QEUH-CC591F/2020\|EPI_ISL_760514\|20 | .................... | .................... | ..................... | .................... | .................... | .................... |
| AB | 7945 | hCoV-19/England/QEUH-CC7AB0/2020\|EPI_ISL_760517\|20 | .................... | .................... | ..................... | .................... | .................... | .................... |
| AB | 7946 | hCoV-19/England/QEUH-CC3B57/2020\|EPI_ISL_760519\|20 | .................... | .................... | ..................... | .................... | .................... | .................... |
| AB | 7947 | hCoV-19/England/ALDP-CBE375/2020\|EPI_ISL_760521\|20 | .................... | .................... | ..................... | .................... | .................... | .................... |
| AB | 7948 | hCoV-19/England/QEUH-CC2341/2020\|EPI_ISL_760525\|20 | .................... | .................... | ..................... | .................... | .................... | .................... |
| AB | 7949 | hCoV-19/England/ALDP-CBDCE4/2020\|EPI_ISL_760531\|20 | .................... | .................... | ..................... | .................... | .................... | .................... |
| AB | 7950 | hCoV-19/England/QEUH-CC7B44/2020\|EPI_ISL_760533\|20 | .................... | .................... | ..................... | .................... | ......T............. | .................... |
| AB | 7951 | hCoV-19/England/ALDP-CBFE28/2020\|EPI_ISL_760535\|20 | .................... | .................... | ..................... | .................... | .................... | .................... |
| AB | 7952 | hCoV-19/England/QEUH-CC2903/2020\|EPI_ISL_760537\|20 | .................... | .................... | ..................... | .................... | .................... | .................... |
| AB | 7953 | hCoV-19/England/QEUH-CC3577/2020\|EPI_ISL_760539\|20 | .................... | .................... | ..................... | .................... | .................... | .................... |
| AB | 7954 | hCoV-19/England/QEUH-CC3D42/2020\|EPI_ISL_760540\|20 | .................... | .................... | ..................... | .................... | .................... | .................... |
| AB | 7955 | hCoV-19/England/QEUH-CC3304/2020\|EPI_ISL_760541\|20 | .................... | .................... | ..................... | .................... | .................... | .................... |

AB 7956 hCoV-19/England/QEUH-CAEBCB/2020|EPI_ISL_760542|20 .................... .A.................. .................... .................... .................... ...A................
AB 7957 hCoV-19/England/ALDP-CBFD76/2020|EPI_ISL_760544|20 .................... .................... .................... .................... .................... ....................
AB 7958 hCoV-19/England/ALDP-CBDE39/2020|EPI_ISL_760545|20 .................... .................... .................... .................... .................... ....................
AB 7959 hCoV-19/England/ALDP-CBE4CD/2020|EPI_ISL_760546|20 .................... .................... .................... .................... .................... ....................
AB 7960 hCoV-19/England/QEUH-CC2271/2020|EPI_ISL_760548|20 .................... .................... .................... .................... .................... ....................
AB 7961 hCoV-19/England/ALDP-CBE603/2020|EPI_ISL_760549|20 .................... .................... .................... .................... .................... ....................
AB 7962 hCoV-19/England/ALDP-CBE58E/2020|EPI_ISL_760551|20 .................... .................... .................... .................... .................... ....................
AB 7963 hCoV-19/England/QEUH-CC2A2E/2020|EPI_ISL_760553|20 .................... .................... .................... .................... .................... ....................
AB 7964 hCoV-19/England/ALDP-CBECB6/2020|EPI_ISL_760555|20 .................... .................... .................... .................... .................... ....................
AB 7965 hCoV-19/England/ALDP-CC1D9F/2020|EPI_ISL_760557|20 .................... .................... .................... .................... .................... ....................
AB 7966 hCoV-19/England/QEUH-CAF32C/2020|EPI_ISL_760560|20 .................... .................... .................... .................... .....T.............. ....................
AB 7967 hCoV-19/England/ALDP-CBE82B/2020|EPI_ISL_760561|20 .................... .................... .................... .................... .................... ....................
AB 7968 hCoV-19/England/ALDP-CC07FC/2020|EPI_ISL_760562|20 .................... .................... .................... .................... .................... ....................
AB 7969 hCoV-19/England/ALDP-CBF5D8/2020|EPI_ISL_760565|20 .................... .................... .................... .................... .................... ....................
AB 7970 hCoV-19/England/QEUH-CC2C55/2020|EPI_ISL_760571|20 .................... .................... .................... .................... .....T.............. ....................
AB 7971 hCoV-19/England/ALDP-CBF602/2020|EPI_ISL_760572|20 .................... .................... .................... .................... .................... ....................
AB 7972 hCoV-19/England/ALDP-CBE533/2020|EPI_ISL_760574|20 .................... .................... .................... .................... .................... ....................
AB 7973 hCoV-19/England/QEUH-CC3911/2020|EPI_ISL_760575|20 .................... .................... .................... .................... .................... ....................
AB 7974 hCoV-19/England/QEUH-CC31CE/2020|EPI_ISL_760576|20 .................... .................... .................... .................... .................... ....................
AB 7975 hCoV-19/England/QEUH-CC397B/2020|EPI_ISL_760580|20 .................... .................... .................... .................... .................... ....................
AB 7976 hCoV-19/England/QEUH-CC3A1E/2020|EPI_ISL_760584|20 .................... .........T.......... .................... .................... .................... ....................
AB 7977 hCoV-19/England/QEUH-CC47F8/2020|EPI_ISL_760585|20 .................... .................... .................... .................... .................... ....................
AB 7978 hCoV-19/England/ALDP-CC061D/2020|EPI_ISL_760587|20 .................... .................... .................... .................... .................... ....................
AB 7979 hCoV-19/England/QEUH-CC3313/2020|EPI_ISL_760590|20 .................... .................... .................... .................... .................... ....................
AB 7980 hCoV-19/England/ALDP-CBF3FC/2020|EPI_ISL_760591|20 .................... .................... .................... .................... .................... ....................
AB 7981 hCoV-19/England/QEUH-CC3D24/2020|EPI_ISL_760592|20 .................... .................... .................... .................... .................... ....................
AB 7982 hCoV-19/England/QEUH-CC4831/2020|EPI_ISL_760594|20 .................... .................... .................... .................... .................... ....................
AB 7983 hCoV-19/England/ALDP-CBE23C/2020|EPI_ISL_760597|20 .................... .................... .................... .................... .................... ....................
AB 7984 hCoV-19/England/ALDP-CBE2C3/2020|EPI_ISL_760599|20 .................... .................... .................... .................... .................... ....................
AB 7985 hCoV-19/England/ALDP-CB4C92/2020|EPI_ISL_761800|20 .................... .................... .................... .................... .................... ....................
AB 7986 hCoV-19/England/ALDP-CB5BC1/2020|EPI_ISL_761802|20 .................... .................... .................... .................... .....T.............. ....................
AB 7987 hCoV-19/England/ALDP-CB65C2/2020|EPI_ISL_761805|20 .................... .................... .................... .................... .................... ....................
AB 7988 hCoV-19/England/ALDP-CB61EC/2020|EPI_ISL_761806|20 .................... .................... .................... .................... .................... ....................
AB 7989 hCoV-19/England/ALDP-CB3765/2020|EPI_ISL_761808|20 .................... .................... .................... .................... .................... ....................
AB 7990 hCoV-19/England/ALDP-CB500E/2020|EPI_ISL_761809|20 .................... .................... .................... .................... .................... ....................
AB 7991 hCoV-19/England/ALDP-CB488F/2020|EPI_ISL_761810|20 .................... .................... .................... .................... .................... ....................
AB 7992 hCoV-19/England/ALDP-CB55F0/2020|EPI_ISL_761811|20 .................... .................... .................... .................... .................... ....................
AB 7993 hCoV-19/England/ALDP-CB6AB4/2020|EPI_ISL_761812|20 .................... .................... .................... .................... .................... ....................
AB 7994 hCoV-19/England/ALDP-CB5138/2020|EPI_ISL_761813|20 .................... .................... .................... .................... .................... ....................
AB 7995 hCoV-19/England/ALDP-CB6814/2020|EPI_ISL_761814|20 .................... .................... .................... .................... .................... ....................
AB 7996 hCoV-19/England/ALDP-CB4843/2020|EPI_ISL_761815|20 .................... .................... .................... .................... .................... ....................
AB 7997 hCoV-19/England/ALDP-CB6A87/2020|EPI_ISL_761817|20 .................... .................... .................... .................... .................... ....................
AB 7998 hCoV-19/England/ALDP-CB4421/2020|EPI_ISL_761818|20 .................... .................... .................... .................... .................... ....................
AB 7999 hCoV-19/England/ALDP-CB6252/2020|EPI_ISL_761820|20 .................... .................... .................... .................... .................... ....................
AB 8000 hCoV-19/England/ALDP-CB5DE9/2020|EPI_ISL_761821|20 .................... .................... .................... .................... .................... ....................
AB 8001 hCoV-19/England/ALDP-CB503B/2020|EPI_ISL_761822|20 .................... .................... .................... .................... .................... ....................
AB 8002 hCoV-19/England/ALDP-CB66A1/2020|EPI_ISL_761823|20 .................... .................... .................... .................... .................... ....................
AB 8003 hCoV-19/England/ALDP-CB6999/2020|EPI_ISL_761825|20 .................... .................... .................... .................... .....T.............. ....................
AB 8004 hCoV-19/England/ALDP-CB6674/2020|EPI_ISL_761826|20 .................... .................... .................... .................... .................... ....................
AB 8005 hCoV-19/England/ALDP-CB4F99/2020|EPI_ISL_761828|20 .................... .................... .................... .................... .................... ....................
AB 8006 hCoV-19/England/ALDP-CB6AE1/2020|EPI_ISL_761829|20 .................... .................... .................... .................... .................... ....................
AB 8007 hCoV-19/England/ALDP-CB5587/2020|EPI_ISL_761830|20 .................... .................... .................... .................... .................... ....................
AB 8008 hCoV-19/England/ALDP-CB4F02/2020|EPI_ISL_761831|20 .................... .................... .................... .................... .................... ....................
AB 8009 hCoV-19/England/ALDP-CB6595/2020|EPI_ISL_761832|20 .................... .................... .................... .................... .................... ....................
AB 8010 hCoV-19/England/ALDP-CB5930/2020|EPI_ISL_761833|20 .................... .................... .................... .................... .................... ....................
AB 8011 hCoV-19/England/ALDP-CB3A7B/2020|EPI_ISL_761837|20 .................... .................... .................... .................... .................... ....................
AB 8012 hCoV-19/England/ALDP-CB636E/2020|EPI_ISL_761838|20 .................... .................... .................... .................... .................... ....................
AB 8013 hCoV-19/England/ALDP-CB5D8F/2020|EPI_ISL_761839|20 .................... .................... .................... .................... .................... ....................
AB 8014 hCoV-19/England/ALDP-CB58AC/2020|EPI_ISL_761840|20 .................... .................... .................... .................... .................... ....................
AB 8015 hCoV-19/England/ALDP-CB5217/2020|EPI_ISL_761841|20 .................... .................... .................... .................... .................... ....................
AB 8016 hCoV-19/England/ALDP-CB53C9/2020|EPI_ISL_761843|20 .................... .................... .................... .................... .................... ....................
AB 8017 hCoV-19/England/ALDP-CB5CCE/2020|EPI_ISL_761844|20 .................... .................... .................... .................... .................... ....................
AB 8018 hCoV-19/England/ALDP-CB4FB7/2020|EPI_ISL_761845|20 .................... .................... .................... .................... .................... ....................
AB 8019 hCoV-19/England/ALDP-CB635F/2020|EPI_ISL_761846|20 .................... .................... .................... .................... .................... ....................
AB 8020 hCoV-19/England/ALDP-CB50A4/2020|EPI_ISL_761849|20 .................... .................... .................... .................... ...T................ ....................
AB 8021 hCoV-19/England/ALDP-CB66FC/2020|EPI_ISL_761850|20 .................... .................... .................... .................... .................... ....................
AB 8022 hCoV-19/England/ALDP-CB589D/2020|EPI_ISL_761852|20 .................... .................... .................... .................... .................... ....................
AB 8023 hCoV-19/England/ALDP-CB65FF/2020|EPI_ISL_761854|20 .................... .................... .................... .................... .................... ....................
AB 8024 hCoV-19/England/ALDP-CB4870/2020|EPI_ISL_761855|20 .................... .................... .................... .................... .................... ....................
AB 8025 hCoV-19/England/ALDP-CB5BA3/2020|EPI_ISL_761856|20 .................... .................... .................... .................... .................... ....................
AB 8026 hCoV-19/England/ALDP-CB66CF/2020|EPI_ISL_761857|20 .................... .................... .................... .................... .................... ....................
AB 8027 hCoV-19/England/ALDP-CB4EF6/2020|EPI_ISL_761859|20 .................... .................... .................... .................... .................... ....................
AB 8028 hCoV-19/England/ALDP-CB52AE/2020|EPI_ISL_761860|20 .................... .................... .................... .................... .................... ....................
AB 8029 hCoV-19/England/ALDP-CB560C/2020|EPI_ISL_761861|20 .................... .................... .................... .................... .................... ....................
AB 8030 hCoV-19/England/ALDP-CB66B0/2020|EPI_ISL_761864|20 .................... .................... .................... .................... .................... ....................
AB 8031 hCoV-19/England/ALDP-CB693F/2020|EPI_ISL_761867|20 .................... .................... .................... .................... .................... ....................
AB 8032 hCoV-19/England/ALDP-CB5262/2020|EPI_ISL_761869|20 .................... .................... .................... .................... .................... ....................
AB 8033 hCoV-19/England/ALDP-CB51CF/2020|EPI_ISL_761872|20 .................... .................... .................... .................... .................... ....................
AB 8034 hCoV-19/England/ALDP-CB5147/2020|EPI_ISL_761873|20 .................... .................... .................... .................... .................... ....................
AB 8035 hCoV-19/England/ALDP-CB60FE/2020|EPI_ISL_761874|20 .................... .................... .................... .................... .................... ....................

| | | | | | | | |
|---|---|---|---|---|---|---|---|
| AB | 8036 hCoV-19/England/ALDP-CB58F7/2020\|EPI_ISL_761876\|20 | ···················· | ···················· | ···················· | ···················· | ···················· | ···················· |
| AB | 8037 hCoV-19/England/ALDP-CB6322/2020\|EPI_ISL_761877\|20 | ···················· | ···················· | ···················· | ···················· | ···················· | ···················· |
| AB | 8038 hCoV-19/England/ALDP-CB598B/2020\|EPI_ISL_761878\|20 | ···················· | ···················· | ···················· | ···················· | ···················· | ···A················ |
| AB | 8039 hCoV-19/England/ALDP-CB4CA1/2020\|EPI_ISL_761879\|20 | ···················· | ···················· | ···················· | ···················· | ······T············· | ···················· |
| AB | 8040 hCoV-19/England/ALDP-CB4C83/2020\|EPI_ISL_761884\|20 | ···················· | ···················· | ···················· | ···················· | ···················· | ···················· |
| AB | 8041 hCoV-19/England/ALDP-CB445E/2020\|EPI_ISL_761885\|20 | ···················· | ···················· | ···················· | ···················· | ···················· | ···················· |
| AB | 8042 hCoV-19/England/ALDP-CB6146/2020\|EPI_ISL_761886\|20 | ···················· | ···················· | ···················· | ···················· | ···················· | ···················· |
| AB | 8043 hCoV-19/England/ALDP-CB501D/2020\|EPI_ISL_761888\|20 | ···················· | ···················· | ···················· | ···················· | ···················· | ···················· |
| AB | 8044 hCoV-19/England/ALDP-CB4F5D/2020\|EPI_ISL_761889\|20 | ···················· | ···················· | ···················· | ···················· | ···················· | ···A················ |
| AB | 8045 hCoV-19/England/ALDP-CB69C6/2020\|EPI_ISL_761890\|20 | ···················· | ···················· | ···················· | ···················· | ······T············· | ···················· |
| AB | 8046 hCoV-19/Wales/ALDP-CB588E/2020\|EPI_ISL_761892\|2020 | ···················· | ···················· | ···················· | ···················· | ···················· | ···················· |
| AB | 8047 hCoV-19/England/ALDP-CB5077/2020\|EPI_ISL_761893\|20 | ···················· | ···················· | ···················· | ···················· | ···················· | ···················· |
| AB | 8048 hCoV-19/England/ALDP-CB3774/2020\|EPI_ISL_761894\|20 | ···················· | ···················· | ···················· | ···················· | ···················· | ···················· |
| AB | 8049 hCoV-19/England/ALDP-CB4C56/2020\|EPI_ISL_761896\|20 | ···················· | ···················· | ···················· | ···················· | ···················· | ···················· |
| AB | 8050 hCoV-19/England/ALDP-CB3729/2020\|EPI_ISL_761897\|20 | ···················· | ···················· | ···················· | ···················· | ···················· | ···················· |
| AB | 8051 hCoV-19/England/ALDP-CB5A97/2020\|EPI_ISL_761898\|20 | ···················· | ···················· | ···················· | ···················· | ···················· | ···················· |
| AB | 8052 hCoV-19/England/ALDP-CB55B4/2020\|EPI_ISL_761899\|20 | ···················· | ···················· | ···················· | ···················· | ···················· | ···················· |
| AB | 8053 hCoV-19/USA/TX-DSHS-2024/2020\|EPI_ISL_766686\|2020- | ···················· | ·············A······ | ···················· | ···················· | ···················· | ···················· |
| AB | 8054 hCoV-19/Sweden/20-53878/2020\|EPI_ISL_766724\|2020-1 | ···················· | ···················· | ···················· | ···················· | ···················· | ···················· |
| AB | 8055 hCoV-19/Sweden/20-53909/2020\|EPI_ISL_766725\|2020-1 | ···················· | ···················· | ···················· | ···················· | ···················· | ···················· |
| AB | 8056 hCoV-19/Sweden/20-53912/2020\|EPI_ISL_766726\|2020-1 | ···················· | ···················· | ···················· | ···················· | ···················· | ···················· |
| AB | 8057 hCoV-19/Sweden/20-53913/2020\|EPI_ISL_766727\|2020-1 | ···················· | ···················· | ···················· | ···················· | ···················· | ···················· |
| AB | 8058 hCoV-19/Sweden/20-53915/2020\|EPI_ISL_766728\|2020-1 | ···················· | ···················· | ···················· | ···················· | ···················· | ···················· |
| AB | 8059 hCoV-19/Sweden/20-53916/2020\|EPI_ISL_766729\|2020-1 | ···················· | ···················· | ···················· | ···················· | ···················· | ···················· |
| AB | 8060 hCoV-19/Sweden/20-53925/2020\|EPI_ISL_766730\|2020-1 | ···················· | ···················· | ···················· | ···················· | ···················· | ···················· |
| AB | 8061 hCoV-19/USA/CO-CDPHE-2100015138/2020\|EPI_ISL_77122 | ···················· | ···················· | ···················· | ···················· | ···················· | ···················· |
| AB | 8062 hCoV-19/USA/CO-CDPHE-2100030884/2020\|EPI_ISL_77122 | ···················· | ·A·················· | ···················· | ···················· | ···················· | ···················· |
| AB | 8063 hCoV-19/USA/CO-CDPHE-2004230036/2020\|EPI_ISL_77122 | ···················· | ···················· | ···················· | ···················· | ···················· | ···················· |
| AB | 8064 hCoV-19/USA/CO-CDPHE-2004250430/2020\|EPI_ISL_77122 | ···················· | ···················· | ···················· | ···················· | ···················· | ···················· |
| AB | 8065 hCoV-19/USA/CO-CDPHE-2005060689/2020\|EPI_ISL_77122 | ···················· | ···················· | ···················· | ···················· | ···················· | ···················· |
| AB | 8066 hCoV-19/USA/CO-CDPHE-2005060754/2020\|EPI_ISL_77122 | ···················· | ···················· | ···················· | ···················· | ···················· | ···················· |
| AB | 8067 hCoV-19/USA/CO-CDPHE-2005060769/2020\|EPI_ISL_77122 | ···················· | ···················· | ···················· | ···················· | ···················· | ···················· |
| AB | 8068 hCoV-19/USA/CO-CDPHE-2007082647/2020\|EPI_ISL_77122 | ···················· | ···················· | ···················· | ···················· | ···················· | ···················· |
| AB | 8069 hCoV-19/USA/CO-CDPHE-2007151943/2020\|EPI_ISL_77123 | ···················· | ···················· | ···················· | ···················· | ···················· | ···················· |
| AB | 8070 hCoV-19/USA/CO-CDPHE-2007152397/2020\|EPI_ISL_77123 | ···················· | ···················· | ···················· | ···················· | ···················· | ···················· |
| AB | 8071 hCoV-19/USA/CO-CDPHE-2007233151/2020\|EPI_ISL_77123 | ···················· | ···················· | ···················· | ···················· | ···················· | ···················· |
| AB | 8072 hCoV-19/USA/CO-CDPHE-2007233534/2020\|EPI_ISL_77123 | ···················· | ···················· | ···················· | ···················· | ···················· | ···················· |
| AB | 8073 hCoV-19/USA/CO-CDPHE-2010230051/2020\|EPI_ISL_77123 | ···················· | ···················· | ···················· | ···················· | ···················· | ···················· |
| AB | 8075 hCoV-19/USA/CO-CDPHE-2011020172/2020\|EPI_ISL_77123 | ···················· | ···················· | ···················· | ···················· | ···················· | ···················· |
| AB | 8076 hCoV-19/USA/CO-CDPHE-2011020202/2020\|EPI_ISL_77123 | ···················· | ···················· | ···················· | ···················· | ···················· | ···················· |
| AB | 8077 hCoV-19/USA/CO-CDPHE-2011020245/2020\|EPI_ISL_77123 | ···················· | ···················· | ···················· | ···················· | ···················· | ···················· |
| AB | 8078 hCoV-19/USA/CO-CDPHE-2011020252/2020\|EPI_ISL_77123 | ···················· | ···················· | ···················· | ···················· | ···················· | ···················· |
| AB | 8079 hCoV-19/USA/CO-CDPHE-2011020263/2020\|EPI_ISL_77124 | ···················· | ···················· | ···················· | ···················· | ···················· | ···················· |
| AB | 8080 hCoV-19/USA/CO-CDPHE-2011020274/2020\|EPI_ISL_77124 | ···················· | ···················· | ···················· | ···················· | ···················· | ···················· |
| AB | 8081 hCoV-19/USA/CO-CDPHE-2011020311/2020\|EPI_ISL_77124 | ···················· | ···················· | ···················· | ···················· | ···················· | ···················· |
| AB | 8082 hCoV-19/USA/CO-CDPHE-2011020331/2020\|EPI_ISL_77124 | ···················· | ···················· | ···················· | ···················· | ···················· | ···················· |
| AB | 8083 hCoV-19/USA/CO-CDPHE-2011020335/2020\|EPI_ISL_77124 | ···················· | ···················· | ···················· | ···················· | ···················· | ···················· |
| AB | 8085 hCoV-19/USA/CO-CDPHE-2011113930/2020\|EPI_ISL_77124 | ···················· | ···················· | ···················· | ···················· | ···················· | ···················· |
| AB | 8086 hCoV-19/USA/CO-CDPHE-2011162142/2020\|EPI_ISL_77124 | ···················· | ···················· | ···················· | ···················· | ···················· | ···················· |
| AB | 8087 hCoV-19/USA/CO-CDPHE-2011165251/2020\|EPI_ISL_77124 | ···················· | ···················· | ···················· | ···················· | ···················· | ···················· |
| AB | 8088 hCoV-19/USA/CO-CDPHE-2011193417/2020\|EPI_ISL_77124 | ···················· | ···················· | ···················· | ···················· | ···················· | ···················· |
| AB | 8089 hCoV-19/USA/CO-CDPHE-2011193669/2020\|EPI_ISL_77125 | ···················· | ·A·················· | ···················· | ···················· | ···················· | ···················· |
| AB | 8090 hCoV-19/USA/CO-CDPHE-2011193707/2020\|EPI_ISL_77125 | ···················· | ···················· | ···················· | ···················· | ···················· | ···················· |
| AB | 8091 hCoV-19/USA/CO-CDPHE-2011193717/2020\|EPI_ISL_77125 | ···················· | ···················· | ···················· | ···················· | ···················· | ···················· |
| AB | 8092 hCoV-19/USA/CO-CDPHE-2011197332/2020\|EPI_ISL_77125 | ···················· | ···················· | ···················· | ···················· | ···················· | ···················· |
| AB | 8093 hCoV-19/USA/CO-CDPHE-2011197416/2020\|EPI_ISL_77125 | ···················· | ···················· | ···················· | ···················· | ···················· | ···················· |
| AB | 8094 hCoV-19/USA/CO-CDPHE-2011199375/2020\|EPI_ISL_77125 | ···················· | ···················· | ···················· | ···················· | ···················· | ···················· |
| AB | 8095 hCoV-19/USA/CO-CDPHE-2011200460/2020\|EPI_ISL_77125 | ···················· | ···················· | ···················· | ···················· | ···················· | ···················· |
| AB | 8096 hCoV-19/USA/CO-CDPHE-2011200477/2020\|EPI_ISL_77125 | ···················· | ···················· | ···················· | ···················· | ···················· | ···················· |
| AB | 8097 hCoV-19/USA/CO-CDPHE-2011200501/2020\|EPI_ISL_77125 | ···················· | ·A·················· | ···················· | ···················· | ···················· | ···················· |
| AB | 8098 hCoV-19/USA/CO-CDPHE-2011200524/2020\|EPI_ISL_77125 | ···················· | ···················· | ···················· | ···················· | ···················· | ···················· |
| AB | 8099 hCoV-19/USA/CO-CDPHE-2011200553/2020\|EPI_ISL_77126 | ···················· | ···················· | ···················· | ···················· | ···················· | ···················· |
| AB | 8100 hCoV-19/USA/CO-CDPHE-2011200641/2020\|EPI_ISL_77126 | ···················· | ···················· | ···················· | ···················· | ···················· | ···················· |
| AB | 8101 hCoV-19/USA/CO-CDPHE-2011201244/2020\|EPI_ISL_77126 | ···················· | ·A·················· | ···················· | ···················· | ···················· | ···················· |
| AB | 8102 hCoV-19/USA/CO-CDPHE-2011210222/2020\|EPI_ISL_77126 | ···················· | ···················· | ···················· | ···················· | ···················· | ···················· |
| AB | 8103 hCoV-19/USA/CO-CDPHE-2011210229/2020\|EPI_ISL_77126 | ···················· | ···················· | ···················· | ···················· | ···················· | ···················· |
| AB | 8104 hCoV-19/USA/CO-CDPHE-2011210235/2020\|EPI_ISL_77126 | ···················· | ···················· | ···················· | ···················· | ···················· | ···················· |
| AB | 8105 hCoV-19/USA/CO-CDPHE-2011210245/2020\|EPI_ISL_77126 | ···················· | ···················· | ···················· | ···················· | ···················· | ···················· |
| AB | 8106 hCoV-19/USA/CO-CDPHE-2011210287/2020\|EPI_ISL_77126 | ···················· | ···················· | ···················· | ···················· | ···················· | ···················· |
| AB | 8107 hCoV-19/USA/CO-CDPHE-2011210294/2020\|EPI_ISL_77126 | ···················· | ···················· | ···················· | ···················· | ···················· | ···················· |
| AB | 8108 hCoV-19/USA/CO-CDPHE-2011210297/2020\|EPI_ISL_77126 | ···················· | ···················· | ···················· | ···················· | ···················· | ···················· |
| AB | 8109 hCoV-19/USA/CO-CDPHE-2011210301/2020\|EPI_ISL_77127 | ···················· | ···················· | ···················· | ···················· | ···················· | ···················· |
| AB | 8110 hCoV-19/USA/CO-CDPHE-2011210336/2020\|EPI_ISL_77127 | ···················· | ···················· | ···················· | ···················· | ···················· | ···················· |
| AB | 8111 hCoV-19/USA/CO-CDPHE-2011210340/2020\|EPI_ISL_77127 | ···················· | ···················· | ···················· | ···················· | ···················· | ···················· |
| AB | 8112 hCoV-19/USA/CO-CDPHE-2011210342/2020\|EPI_ISL_77127 | ···················· | ···················· | ···················· | ···················· | ···················· | ···················· |
| AB | 8113 hCoV-19/USA/CO-CDPHE-2011210344/2020\|EPI_ISL_77127 | ···················· | ···················· | ···················· | ···················· | ···················· | ···················· |
| AB | 8114 hCoV-19/USA/CO-CDPHE-2011210349/2020\|EPI_ISL_77127 | ···················· | ···················· | ···················· | ···················· | ···················· | ···················· |
| AB | 8115 hCoV-19/USA/CO-CDPHE-2011210352/2020\|EPI_ISL_77127 | ···················· | ···················· | ···················· | ···················· | ···················· | ···················· |
| AB | 8116 hCoV-19/USA/CO-CDPHE-2011210368/2020\|EPI_ISL_77127 | ···················· | ···················· | ···················· | ···················· | ···················· | ···················· |
| AB | 8117 hCoV-19/Germany/HH-hpi-p1571/2020\|EPI_ISL_776050\|2 | ···················· | ···················· | ···················· | ···················· | ···················· | ···················· |

| | | | | | | | |
|---|---|---|---|---|---|---|---|
| AB | 8118 hCoV-19/Germany/HH-hpi-p1572/2020\|EPI_ISL_776051\|2 | ···················· | ···················· | ···················· | ···················· | ···················· | ···················· |
| AB | 8119 hCoV-19/Germany/HH-hpi-p1574/2020\|EPI_ISL_776052\|2 | ···················· | ···················· | ···················· | ···················· | ···················· | ···················· |
| AB | 8120 hCoV-19/Germany/HH-hpi-p1575/2020\|EPI_ISL_776053\|2 | ···················· | ···················· | ···················· | ···················· | ···················· | ···················· |
| AB | 8121 hCoV-19/Germany/HH-hpi-p1576/2020\|EPI_ISL_776054\|2 | ···················· | ···················· | ···················· | ···················· | ···················· | ···················· |
| AB | 8122 hCoV-19/Germany/HH-hpi-p1577/2020\|EPI_ISL_776055\|2 | ···················· | ···················· | ···················· | ···················· | ···················· | ···················· |
| AB | 8124 hCoV-19/Germany/HH-hpi-p1582/2020\|EPI_ISL_776057\|2 | ···················· | ···················· | ···················· | ···················· | ···················· | ···················· |
| AB | 8125 hCoV-19/Germany/HH-hpi-p1583/2020\|EPI_ISL_776058\|2 | ···················· | ···················· | ···················· | ···················· | ···················· | ···················· |
| AB | 8126 hCoV-19/Germany/HH-hpi-p1584/2020\|EPI_ISL_776059\|2 | ···················· | ···················· | ···················· | ···················· | ···················· | ···················· |
| AB | 8127 hCoV-19/Germany/HH-hpi-p1588/2020\|EPI_ISL_776060\|2 | ···················· | ···················· | ···················· | ···················· | ···················· | ···················· |
| AB | 8128 hCoV-19/Germany/HH-hpi-p1589/2020\|EPI_ISL_776061\|2 | ···················· | ···················· | ···················· | ···················· | ···················· | ···················· |
| AB | 8129 hCoV-19/Germany/HH-hpi-p1590/2020\|EPI_ISL_776062\|2 | ···················· | ···················· | ···················· | ···················· | ···················· | ···················· |
| AB | 8130 hCoV-19/Germany/HH-hpi-p1591/2020\|EPI_ISL_776063\|2 | ···················· | ···················· | ···················· | ···················· | ···················· | ···················· |
| AB | 8131 hCoV-19/Germany/HH-hpi-p1597/2020\|EPI_ISL_776064\|2 | ···················· | ···················· | ···················· | ···················· | ···················· | ···················· |
| AB | 8132 hCoV-19/Germany/HH-hpi-p1598/2020\|EPI_ISL_776065\|2 | ···················· | ···················· | ···················· | ···················· | ···················· | ···················· |
| AB | 8133 hCoV-19/Germany/HH-hpi-p1606/2020\|EPI_ISL_776066\|2 | ···················· | ···················· | ···················· | ···················· | ···················· | ···················· |
| AB | 8134 hCoV-19/Germany/HH-hpi-p1611/2020\|EPI_ISL_776067\|2 | ···················· | ···················· | ···················· | ···················· | ···················· | ···················· |
| AB | 8135 hCoV-19/Germany/HH-hpi-p1613/2020\|EPI_ISL_776068\|2 | ···················· | ···················· | ···················· | ···················· | ···················· | ···················· |
| AB | 8136 hCoV-19/Germany/HH-hpi-p1615/2020\|EPI_ISL_776069\|2 | ···················· | ···················· | ···················· | ···················· | ···················· | ···················· |
| AB | 8137 hCoV-19/Germany/HH-hpi-p1616/2020\|EPI_ISL_776070\|2 | ···················· | ···················· | ···················· | ···················· | ···················· | ···················· |
| AB | 8138 hCoV-19/Germany/HH-hpi-p1617/2020\|EPI_ISL_776071\|2 | ···················· | ···················· | ···················· | ···················· | ···················· | ···················· |
| AB | 8139 hCoV-19/Germany/HH-hpi-p1619/2020\|EPI_ISL_776072\|2 | ···················· | ···················· | ···················· | ···················· | ···················· | ···················· |
| AB | 8140 hCoV-19/Germany/HH-hpi-p1620/2020\|EPI_ISL_776073\|2 | ···················· | ···················· | ···················· | ···················· | ···················· | ···················· |
| AB | 8141 hCoV-19/Germany/HH-hpi-p1622/2020\|EPI_ISL_776074\|2 | ···················· | ···················· | ···················· | ···················· | ···················· | ···················· |
| AB | 8142 hCoV-19/Germany/HH-hpi-p1623/2020\|EPI_ISL_776075\|2 | ···················· | ···················· | ···················· | ···················· | ···················· | ···················· |
| AB | 8143 hCoV-19/Germany/HH-hpi-p1626/2020\|EPI_ISL_776076\|2 | ···················· | ···················· | ···················· | ···················· | ···················· | ···················· |
| AB | 8144 hCoV-19/Germany/HH-hpi-p1628/2020\|EPI_ISL_776077\|2 | ···················· | ···················· | ···················· | ···················· | ···················· | ···················· |
| AB | 8145 hCoV-19/Germany/HH-hpi-p1629/2020\|EPI_ISL_776078\|2 | ···················· | ···················· | ···················· | ···················· | ···················· | ···················· |
| AB | 8146 hCoV-19/Germany/HH-hpi-p1639/2020\|EPI_ISL_776079\|2 | ···················· | ···················· | ···················· | ···················· | ···················· | ···················· |
| AB | 8147 hCoV-19/Germany/HH-hpi-p1643/2020\|EPI_ISL_776080\|2 | ···················· | ···················· | ···················· | ···················· | ···················· | ···················· |
| AB | 8148 hCoV-19/Germany/HH-hpi-p1647/2020\|EPI_ISL_776081\|2 | ···················· | ···················· | ···················· | ···················· | ···················· | ···················· |
| AB | 8149 hCoV-19/Germany/HH-hpi-p1648/2020\|EPI_ISL_776082\|2 | ···················· | ···················· | ···················· | ···················· | ···················· | ···················· |
| AB | 8150 hCoV-19/Germany/HH-hpi-p1652/2020\|EPI_ISL_776083\|2 | ···················· | ···················· | ···················· | ···················· | ···················· | ···················· |
| AB | 8151 hCoV-19/Germany/HH-hpi-p1658/2020\|EPI_ISL_776084\|2 | ···················· | ···················· | ···················· | ···················· | ···················· | ···················· |
| AB | 8152 hCoV-19/Germany/HH-hpi-p1659/2020\|EPI_ISL_776085\|2 | ···················· | ···················· | ···················· | ···················· | ···················· | ···················· |
| AB | 8153 hCoV-19/Germany/HH-hpi-p1661/2020\|EPI_ISL_776086\|2 | ···················· | ···················· | ···················· | ···················· | ···················· | ···················· |
| AB | 8154 hCoV-19/Germany/HH-hpi-p1665/2020\|EPI_ISL_776087\|2 | ···················· | ···················· | ···················· | ···················· | ···················· | ···················· |
| AB | 8155 hCoV-19/Germany/HH-hpi-p1666/2020\|EPI_ISL_776088\|2 | ···················· | ···················· | ···················· | ···················· | ···················· | ···················· |
| AB | 8156 hCoV-19/Germany/HH-hpi-p1669/2020\|EPI_ISL_776089\|2 | ···················· | ···················· | ···················· | ···················· | ···················· | ···················· |
| AB | 8157 hCoV-19/Germany/HH-hpi-p1670/2020\|EPI_ISL_776090\|2 | ···················· | ···················· | ···················· | ···················· | ···················· | ···················· |
| AB | 8158 hCoV-19/Germany/HH-hpi-p1675/2020\|EPI_ISL_776091\|2 | ···················· | ···················· | ···················· | ···················· | ···················· | ···················· |
| AB | 8159 hCoV-19/Germany/HH-hpi-p1677/2020\|EPI_ISL_776092\|2 | ···················· | ···················· | ···················· | ···················· | ···················· | ···················· |
| AB | 8160 hCoV-19/Germany/HH-hpi-p1678/2020\|EPI_ISL_776093\|2 | ···················· | ···················· | ···················· | ···················· | ···················· | ···················· |
| AB | 8161 hCoV-19/Germany/HH-hpi-p1680/2020\|EPI_ISL_776094\|2 | ···················· | ···················· | ···················· | ···················· | ···················· | ···················· |
| AB | 8162 hCoV-19/Germany/HH-hpi-p1684/2020\|EPI_ISL_776095\|2 | ···················· | ···················· | ···················· | ···················· | ···················· | ···················· |
| AB | 8163 hCoV-19/Germany/HH-hpi-p1686/2020\|EPI_ISL_776096\|2 | ···················· | ···················· | ···················· | ···················· | ···················· | ···················· |
| AB | 8164 hCoV-19/Germany/HH-hpi-p1687/2020\|EPI_ISL_776097\|2 | ···················· | ···················· | ···················· | ···················· | ···················· | ···················· |
| AB | 8165 hCoV-19/Germany/HH-hpi-p1688/2020\|EPI_ISL_776098\|2 | ···················· | ···················· | ···················· | ···················· | ···················· | ···················· |
| AB | 8166 hCoV-19/Germany/HH-hpi-p1689/2020\|EPI_ISL_776099\|2 | ···················· | ···················· | ···················· | ···················· | ···················· | ···················· |
| AB | 8167 hCoV-19/England/QEUH-C93895/2020\|EPI_ISL_777322\|20 | ···················· | ···················· | ···················· | ···················· | ···················· | ···················· |
| AB | 8168 hCoV-19/England/QEUH-C94885/2020\|EPI_ISL_777324\|20 | ···················· | ···················· | ···················· | ···················· | ·····T·············· | ···················· |
| AB | 8169 hCoV-19/England/QEUH-C939CF/2020\|EPI_ISL_777325\|20 | ···················· | ···················· | ···················· | ···················· | ···················· | ···················· |
| AB | 8170 hCoV-19/England/ALDP-C8C02E/2020\|EPI_ISL_777326\|20 | ···················· | ···················· | ···················· | ···················· | ···················· | ···················· |
| AB | 8171 hCoV-19/England/ALDP-C8DE32/2020\|EPI_ISL_777380\|20 | ···················· | ···················· | ···················· | ···················· | ···················· | ···················· |
| AB | 8172 hCoV-19/England/CAMC-C98E52/2020\|EPI_ISL_777381\|20 | ···················· | ···················· | ···················· | ···················· | ···················· | ···················· |
| AB | 8173 hCoV-19/England/CAMC-C99088/2020\|EPI_ISL_777387\|20 | ···················· | ·A·················· | ···················· | ···················· | ···················· | ···················· |
| AB | 8174 hCoV-19/England/CAMC-C98906/2020\|EPI_ISL_777390\|20 | ···················· | ···················· | ···················· | ···················· | ···················· | ···················· |
| AB | 8175 hCoV-19/England/ALDP-C8E693/2020\|EPI_ISL_777394\|20 | ···················· | ···················· | ···················· | ···················· | ···················· | ···················· |
| AB | 8176 hCoV-19/England/QEUH-C93E1B/2020\|EPI_ISL_777395\|20 | ···················· | ···················· | ···················· | ···················· | ···················· | ···················· |
| AB | 8177 hCoV-19/England/QEUH-C93F54/2020\|EPI_ISL_777397\|20 | ···················· | ···················· | ···················· | ···················· | ···················· | ···················· |
| AB | 8178 hCoV-19/Italy/CAM-TIGEM-654/2020\|EPI_ISL_778651\|20 | ···················· | ···················· | ···················· | ···················· | ···················· | ···················· |
| AB | 8179 hCoV-19/Italy/CAM-TIGEM-611/2020\|EPI_ISL_778652\|20 | ···················· | ···················· | ···················· | ···················· | ···················· | ···················· |
| AB | 8180 hCoV-19/Italy/CAM-TIGEM-603/2020\|EPI_ISL_778653\|20 | ···················· | ···················· | ···················· | ···················· | ···················· | ···················· |
| AB | 8181 hCoV-19/Italy/CAM-TIGEM-634/2020\|EPI_ISL_778654\|20 | ···················· | ···················· | ···················· | ···················· | ···················· | ···················· |
| AB | 8182 hCoV-19/Italy/CAM-TIGEM-613/2020\|EPI_ISL_778655\|20 | ···················· | ···················· | ···················· | ···················· | ···················· | ···················· |
| AB | 8183 hCoV-19/Italy/CAM-TIGEM-614/2020\|EPI_ISL_778656\|20 | ···················· | ···················· | ···················· | ···················· | ···················· | ···················· |
| AB | 8184 hCoV-19/Italy/CAM-TIGEM-585/2020\|EPI_ISL_778657\|20 | ···················· | ···················· | ···················· | ···················· | ···················· | ···················· |
| AB | 8185 hCoV-19/Italy/CAM-TIGEM-579/2020\|EPI_ISL_778658\|20 | ···················· | ···················· | ···················· | ···················· | ···················· | ···················· |
| AB | 8186 hCoV-19/Italy/CAM-TIGEM-419/2020\|EPI_ISL_778714\|20 | ···················· | ···················· | ···················· | ···················· | ·····T·············· | ···················· |
| AB | 8187 hCoV-19/Italy/CAM-TIGEM-404/2020\|EPI_ISL_778715\|20 | ···················· | ···················· | ···················· | ···················· | ·····T·············· | ···················· |
| AB | 8188 hCoV-19/Italy/CAM-TIGEM-399/2020\|EPI_ISL_778716\|20 | ···················· | ···················· | ···················· | ···················· | ·····T·············· | ···················· |
| AB | 8189 hCoV-19/Italy/CAM-TIGEM-408/2020\|EPI_ISL_778717\|20 | ···················· | ···················· | ···················· | ···················· | ···················· | ···················· |
| AB | 8190 hCoV-19/Italy/CAM-TIGEM-406/2020\|EPI_ISL_778719\|20 | ···················· | ···················· | ···················· | ···················· | ·····T·············· | ···················· |
| AB | 8191 hCoV-19/Italy/CAM-TIGEM-553/2020\|EPI_ISL_778723\|20 | ···················· | ···················· | ···················· | ···················· | ···················· | ···················· |
| AB | 8192 hCoV-19/Italy/CAM-TIGEM-548/2020\|EPI_ISL_778724\|20 | ···················· | ···················· | ···················· | ···················· | ···················· | ···················· |
| AB | 8193 hCoV-19/England/ALDP-CB5E40/2020\|EPI_ISL_761900\|20 | ···················· | ···················· | ···················· | ···················· | ···················· | ···················· |
| AB | 8194 hCoV-19/England/ALDP-CB3A4E/2020\|EPI_ISL_761901\|20 | ···················· | ···················· | ···················· | ···················· | ···················· | ···················· |
| AB | 8195 hCoV-19/England/ALDP-CB4403/2020\|EPI_ISL_761903\|20 | ···················· | ···················· | ···················· | ···················· | ···················· | ···················· |
| AB | 8196 hCoV-19/England/ALDP-CB4F8A/2020\|EPI_ISL_761904\|20 | ···················· | ···················· | ···················· | ···················· | ·····T·············· | ···················· |
| AB | 8197 hCoV-19/England/ALDP-CB4BE0/2020\|EPI_ISL_761908\|20 | ···················· | ···················· | ···················· | ···················· | ···················· | ···················· |
| AB | 8198 hCoV-19/England/ALDP-CB37FC/2020\|EPI_ISL_761909\|20 | ···················· | ···················· | ···················· | ···················· | ···················· | ···················· |

| | | | | | | | | |
|---|---|---|---|---|---|---|---|---|
| AB | 8199 hCoV-19/England/SHEF-10C607C/2020\|EPI_ISL_764103\|2 | ···················· | ···················· | ···················· | ···················· | ···················· | ···················· | |
| AB | 8200 hCoV-19/England/SHEF-10C6443/2020\|EPI_ISL_764104\|2 | ···················· | ···················· | ···················· | ···················· | ···················· | ···················· | |
| AB | 8201 hCoV-19/England/SHEF-10C9E42/2020\|EPI_ISL_764105\|2 | ···················· | ···················· | ···················· | ···················· | ···················· | ···················· | |
| AB | 8202 hCoV-19/England/SHEF-10CB39C/2020\|EPI_ISL_764106\|2 | ···················· | ···················· | ···················· | ···················· | ···················· | ···················· | |
| AB | 8203 hCoV-19/England/SHEF-10CFEC4/2020\|EPI_ISL_764107\|2 | ···················· | ···················· | ···················· | ···················· | ···················· | ···················· | |
| AB | 8204 hCoV-19/England/SHEF-10D0C20/2020\|EPI_ISL_764108\|2 | ···················· | ···················· | ···················· | ···················· | ···················· | ···················· | |
| AB | 8205 hCoV-19/England/SHEF-10D38FC/2020\|EPI_ISL_764109\|2 | ···················· | ···················· | ···················· | ···················· | ···················· | ···················· | |
| AB | 8206 hCoV-19/England/SHEF-10D4512/2020\|EPI_ISL_764110\|2 | ···················· | ···················· | ···················· | ···················· | ···················· | ···················· | |
| AB | 8207 hCoV-19/England/SHEF-10D510E/2020\|EPI_ISL_764111\|2 | ···················· | ···················· | ···················· | ···················· | ···················· | ···················· | |
| AB | 8208 hCoV-19/England/SHEF-10D6844/2020\|EPI_ISL_764112\|2 | ···················· | ···················· | ···················· | ···················· | ···················· | ···················· | |
| AB | 8209 hCoV-19/England/NORW-222367/2020\|EPI_ISL_764113\|20 | ···················· | ···················· | ···················· | ···················· | ···················· | ···················· | |
| AB | 8210 hCoV-19/England/NORW-2224A0/2020\|EPI_ISL_764114\|20 | ···················· | ···················· | ···················· | ···················· | ···················· | ···················· | |
| AB | 8211 hCoV-19/England/NORW-2224EC/2020\|EPI_ISL_764115\|20 | ···················· | ···················· | ···················· | ···················· | ···················· | ···················· | |
| AB | 8212 hCoV-19/England/NORW-222886/2020\|EPI_ISL_764116\|20 | ···················· | ···················· | ···················· | ···················· | ···················· | ···················· | |
| AB | 8213 hCoV-19/England/OXON-F409FB/2020\|EPI_ISL_764117\|20 | ···················· | ···················· | ···················· | ···················· | ···················· | ···················· | |
| AB | 8214 hCoV-19/England/PORT-2D55B8/2020\|EPI_ISL_764118\|20 | ···················· | ·A·················· | ···················· | ···················· | ···················· | ··A················· | |
| AB | 8216 hCoV-19/England/PORT-2D5819/2020\|EPI_ISL_764120\|20 | ···················· | ·A·················· | ···················· | ···················· | ···················· | ··A················· | |
| AB | 8217 hCoV-19/England/PORT-2D58CE/2020\|EPI_ISL_764121\|20 | ···················· | ·A·················· | ···················· | ···················· | ···················· | ··A················· | |
| AB | 8218 hCoV-19/England/PORT-2D59DA/2020\|EPI_ISL_764122\|20 | ···················· | ·A·················· | ···················· | ···················· | ···················· | ··A················· | |
| AB | 8219 hCoV-19/England/PORT-2D5A8C/2020\|EPI_ISL_764123\|20 | ···················· | ·A·················· | ···················· | ···················· | ···················· | ··A················· | |
| AB | 8220 hCoV-19/England/PORT-2D5CD1/2020\|EPI_ISL_764124\|20 | ···················· | ·A·················· | ···················· | ···················· | ···················· | ··A················· | |
| AB | 8221 hCoV-19/England/PORT-2D5E44/2020\|EPI_ISL_764125\|20 | ···················· | ·A·················· | ···················· | ···················· | ···················· | ··A················· | |
| AB | 8222 hCoV-19/England/NORW-2228B3/2020\|EPI_ISL_764126\|20 | ···················· | ·A·················· | ···················· | ···················· | ···················· | ··A················· | |
| AB | 8224 hCoV-19/USA/NY-Wadsworth-290335-01/2020\|EPI_ISL_76 | ···················· | ···---------········ | ···················· | ···················· | ···················· | ···················· | 1 |
| AB | 8225 hCoV-19/USA/NY-Wadsworth-290386-01/2020\|EPI_ISL_76 | ···················· | ···················· | ···················· | ···················· | ···················· | ···················· | |
| AB | 8226 hCoV-19/USA/NY-Wadsworth-213974-01/2020\|EPI_ISL_76 | ···················· | ···················· | ···················· | ···················· | ···················· | ···················· | |
| AB | 8227 hCoV-19/USA/NY-Wadsworth-213976-01/2020\|EPI_ISL_76 | ···················· | ···················· | ···················· | ···················· | ···················· | ···················· | |
| AB | 8228 hCoV-19/USA/NY-Wadsworth-213917-01/2020\|EPI_ISL_76 | ···················· | ···················· | ···················· | ···················· | ···················· | ···················· | |
| AB | 8229 hCoV-19/USA/NY-Wadsworth-213736-01/2020\|EPI_ISL_76 | ···················· | ···················· | ···················· | ···················· | ···················· | ···················· | |
| AB | 8230 hCoV-19/Sweden/20-33499/2020\|EPI_ISL_766700\|2020-1 | ···················· | ···················· | ···················· | ···················· | ···················· | ···················· | |
| AB | 8231 hCoV-19/Sweden/20-33500/2020\|EPI_ISL_766701\|2020-1 | ···················· | ···················· | ···················· | ···················· | ···················· | ···················· | |
| AB | 8232 hCoV-19/Sweden/20-33504/2020\|EPI_ISL_766702\|2020-1 | ···················· | ···················· | ···················· | ···················· | ···················· | ···················· | |
| AB | 8233 hCoV-19/Sweden/20-33507/2020\|EPI_ISL_766703\|2020-1 | ···················· | ···················· | ···················· | ···················· | ···················· | ···················· | |
| AB | 8234 hCoV-19/Sweden/20-33508/2020\|EPI_ISL_766704\|2020-1 | ···················· | ···················· | ···················· | ···················· | ···················· | ···················· | |
| AB | 8235 hCoV-19/Sweden/20-33512/2020\|EPI_ISL_766705\|2020-1 | ···················· | ···················· | ···················· | ···················· | ···················· | ···················· | |
| AB | 8236 hCoV-19/Sweden/20-33522/2020\|EPI_ISL_766706\|2020-1 | ···················· | ···················· | ···················· | ···················· | ···················· | ···················· | |
| AB | 8237 hCoV-19/Sweden/20-53769/2020\|EPI_ISL_766707\|2020-1 | ···················· | ···················· | ···················· | ···················· | ···················· | ···················· | |
| AB | 8238 hCoV-19/Sweden/20-13184/2020\|EPI_ISL_766708\|2020-1 | ···················· | ···················· | ···················· | ···················· | ···················· | ···················· | |
| AB | 8239 hCoV-19/Sweden/20-53765/2020\|EPI_ISL_766710\|2020-1 | ···················· | ···················· | ···················· | ···················· | ···················· | ···················· | |
| AB | 8240 hCoV-19/Sweden/20-53768/2020\|EPI_ISL_766711\|2020-1 | ···················· | ···················· | ···················· | ···················· | ···················· | ···················· | |
| AB | 8241 hCoV-19/Sweden/20-53775/2020\|EPI_ISL_766712\|2020-1 | ···················· | ···················· | ···················· | ···················· | ···················· | ···················· | |
| AB | 8242 hCoV-19/Sweden/20-53794/2020\|EPI_ISL_766713\|2020-1 | ···················· | ···················· | ···················· | ···················· | ···················· | ···················· | |
| AB | 8243 hCoV-19/Sweden/20-13161/2020\|EPI_ISL_766714\|2020-1 | ···················· | ···················· | ···················· | ···················· | ···················· | ···················· | |
| AB | 8244 hCoV-19/Sweden/20-13165/2020\|EPI_ISL_766715\|2020-1 | ···················· | ···················· | ···················· | ···················· | ···················· | ···················· | |
| AB | 8245 hCoV-19/Sweden/20-13166/2020\|EPI_ISL_766716\|2020-1 | ···················· | ···················· | ···················· | ···················· | ··C················· | ···················· | |
| AB | 8246 hCoV-19/Sweden/20-13170/2020\|EPI_ISL_766717\|2020-1 | ···················· | ···················· | ···················· | ···················· | ···················· | ···················· | |
| AB | 8247 hCoV-19/Sweden/20-13172/2020\|EPI_ISL_766718\|2020-1 | ···················· | ···················· | ···················· | ···················· | ···················· | ···················· | |
| AB | 8248 hCoV-19/Sweden/20-13176/2020\|EPI_ISL_766719\|2020-1 | ···················· | ···················· | ···················· | ···················· | ···················· | ···················· | |
| AB | 8249 hCoV-19/Sweden/20-13178/2020\|EPI_ISL_766720\|2020-1 | ···················· | ···················· | ···················· | ···················· | ···················· | ···················· | |
| AB | 8250 hCoV-19/Sweden/20-13179/2020\|EPI_ISL_766721\|2020-1 | ···················· | ···················· | ···················· | ···················· | ···················· | ···················· | |
| AB | 8251 hCoV-19/Sweden/20-13180/2020\|EPI_ISL_766722\|2020-1 | ···················· | ···················· | ···················· | ···················· | ···················· | ···················· | |
| AB | 8252 hCoV-19/Sweden/20-13182/2020\|EPI_ISL_766723\|2020-1 | ···················· | ···················· | ···················· | ···················· | ···················· | ···················· | |
| AB | 8253 hCoV-19/USA/TX-DSHS-2008/2020\|EPI_ISL_766762\|2020- | ···················· | ·A·················· | ···················· | ···················· | ···················· | ···················· | |
| AB | 8254 hCoV-19/USA/TX-DSHS-2010/2020\|EPI_ISL_766763\|2020- | ···················· | ·A·················· | ···················· | ···················· | ···················· | ···················· | |
| AB | 8255 hCoV-19/USA/TX-DSHS-2011/2020\|EPI_ISL_766764\|2020- | ···················· | ·A·················· | ···················· | ···················· | ···················· | ···················· | |
| AB | 8256 hCoV-19/USA/TX-DSHS-1971/2020\|EPI_ISL_766765\|2020- | ···················· | ···················· | ···················· | ···················· | ···················· | ···················· | |
| AB | 8257 hCoV-19/USA/TX-DSHS-1979/2020\|EPI_ISL_766766\|2020- | ···················· | ···················· | ···················· | ···················· | ···················· | ···················· | |
| AB | 8258 hCoV-19/USA/TX-DSHS-1981/2020\|EPI_ISL_766767\|2020- | ···················· | ···················· | ···················· | ···················· | ···················· | ···················· | |
| AB | 8259 hCoV-19/USA/TX-DSHS-1982/2020\|EPI_ISL_766768\|2020- | ···················· | ···················· | ···················· | ···················· | ···················· | ···················· | |
| AB | 8260 hCoV-19/USA/TX-DSHS-1984/2020\|EPI_ISL_766769\|2020- | ···················· | ···················· | ···················· | ···················· | ···················· | ···················· | |
| AB | 8261 hCoV-19/USA/TX-DSHS-1987/2020\|EPI_ISL_766770\|2020- | ···················· | ···················· | ···················· | ···················· | ···················· | ···················· | |
| AB | 8262 hCoV-19/USA/TX-DSHS-1992/2020\|EPI_ISL_766771\|2020- | ···················· | ···················· | ···················· | ···················· | ···················· | ···················· | |
| AB | 8263 hCoV-19/USA/TX-DSHS-1995/2020\|EPI_ISL_766772\|2020- | ···················· | ·A·················· | ···················· | ···················· | ···················· | ···················· | |
| AB | 8264 hCoV-19/USA/WY-WPHL-20167881/2020\|EPI_ISL_766775\|2 | ·········T·········· | ···················· | ···················· | ···················· | ···················· | ···················· | |
| AB | 8265 hCoV-19/USA/WY-WPHL-20167931/2020\|EPI_ISL_766776\|2 | ···················· | ···················· | ···················· | ···················· | ···················· | ···················· | |
| AB | 8266 hCoV-19/USA/WY-WPHL-20167940/2020\|EPI_ISL_766777\|2 | ···················· | ···················· | ···················· | ···················· | ···················· | ···················· | |
| AB | 8267 hCoV-19/USA/WY-WPHL-20167949/2020\|EPI_ISL_766778\|2 | ···················· | ···················· | ···················· | ···················· | ···················· | ···················· | |
| AB | 8268 hCoV-19/USA/WY-WPHL-20168020/2020\|EPI_ISL_766779\|2 | ···················· | ···················· | ···················· | ···················· | ···················· | ···················· | |
| AB | 8269 hCoV-19/USA/WY-WPHL-20168039/2020\|EPI_ISL_766780\|2 | ···················· | ···················· | ···················· | ···················· | ···················· | ···················· | |
| AB | 8270 hCoV-19/USA/WY-WPHL-20168040/2020\|EPI_ISL_766781\|2 | ···················· | ···················· | ···················· | ···················· | ···················· | ···················· | |
| AB | 8271 hCoV-19/USA/WY-WPHL-20168068/2020\|EPI_ISL_766782\|2 | ···················· | ···················· | ···················· | ···················· | ···················· | ···················· | |
| AB | 8272 hCoV-19/USA/WY-WPHL-20168086/2020\|EPI_ISL_766783\|2 | ···················· | ···················· | ···················· | ···················· | ···················· | ···················· | |
| AB | 8273 hCoV-19/USA/WY-WPHL-20168100/2020\|EPI_ISL_766784\|2 | ···················· | ·A·················· | ···················· | ···················· | ···················· | ···················· | |
| AB | 8274 hCoV-19/USA/WY-WPHL-20168129/2020\|EPI_ISL_766785\|2 | ···················· | ·A·················· | ···················· | ···················· | ···················· | ···················· | |
| AB | 8275 hCoV-19/USA/WY-WPHL-20168140/2020\|EPI_ISL_766786\|2 | ···················· | ·A·················· | ···················· | ···················· | ···················· | ···················· | |
| AB | 8276 hCoV-19/USA/WY-WPHL-20168141/2020\|EPI_ISL_766787\|2 | ···················· | ···················· | ···················· | ···················· | ···················· | ···················· | |
| AB | 8277 hCoV-19/USA/WY-WPHL-20168166/2020\|EPI_ISL_766788\|2 | ···················· | ·A·················· | ···················· | ···················· | ···················· | ···················· | |
| AB | 8278 hCoV-19/USA/WY-WPHL-20168359/2020\|EPI_ISL_766790\|2 | ···················· | ···················· | ···················· | ···················· | ···················· | ···················· | |
| AB | 8279 hCoV-19/USA/CO-CDPHE-2011210416/2020\|EPI_ISL_77127 | ···················· | ···················· | ···················· | ···················· | ···················· | ···················· | |
| AB | 8280 hCoV-19/USA/CO-CDPHE-2011233599/2020\|EPI_ISL_77127 | ···················· | ···················· | ···················· | ···················· | ···················· | ···················· | |

| | | | | | | | | |
|---|---|---|---|---|---|---|---|---|
| AB | 8281 | hCoV-19/USA/CO-CDPHE-2100000841/2020\|EPI_ISL_77128 | ········T··········· | ·A·················· | ···················· | ···················· | ···················· | ···················· |
| AB | 8282 | hCoV-19/USA/CO-CDPHE-2100000948/2020\|EPI_ISL_77128 | ···················· | ···················· | ···················· | ···················· | ···················· | ···················· |
| AB | 8283 | hCoV-19/USA/CO-CDPHE-2100001027/2020\|EPI_ISL_77128 | ···················· | ·A·················· | ···················· | ···················· | ···················· | ···················· |
| AB | 8284 | hCoV-19/USA/CO-CDPHE-2100001047/2020\|EPI_ISL_77128 | ···················· | ·A·················· | ···················· | ···················· | ···················· | ···················· |
| AB | 8285 | hCoV-19/USA/CO-CDPHE-2100004476/2020\|EPI_ISL_77128 | ···················· | ·A·················· | ···················· | ···················· | ···················· | ···················· |
| AB | 8286 | hCoV-19/USA/CO-CDPHE-2100004497/2020\|EPI_ISL_77128 | ···················· | ···················· | ···················· | ···················· | ···················· | ···················· |
| AB | 8287 | hCoV-19/USA/CO-CDPHE-2100004572/2020\|EPI_ISL_77128 | ···················· | ···················· | ···················· | ···················· | ···················· | ···················· |
| AB | 8288 | hCoV-19/USA/CO-CDPHE-2100005087/2020\|EPI_ISL_77128 | ···················· | ·A·················· | ···················· | ···················· | ···················· | ···················· |
| AB | 8289 | hCoV-19/USA/CO-CDPHE-2100005207/2020\|EPI_ISL_77128 | ···················· | ·A·················· | ···················· | ···················· | ···················· | ···················· |
| AB | 8290 | hCoV-19/USA/CO-CDPHE-2100005226/2020\|EPI_ISL_77128 | ···················· | ·A·················· | ···················· | ···················· | ···················· | ···················· |
| AB | 8291 | hCoV-19/USA/CO-CDPHE-2100005818/2020\|EPI_ISL_77129 | ···················· | ···················· | ···················· | ···················· | ···················· | ···················· |
| AB | 8292 | hCoV-19/USA/CO-CDPHE-2100008783/2020\|EPI_ISL_77129 | ···················· | ···················· | ···················· | ···················· | ···················· | ···················· |
| AB | 8293 | hCoV-19/USA/CO-CDPHE-2100009945/2020\|EPI_ISL_77129 | ···················· | ···················· | ···················· | ···················· | ···················· | ···················· |
| AB | 8295 | hCoV-19/USA/CO-CDPHE-2100010220/2020\|EPI_ISL_77129 | ········T··········· | ···················· | ···················· | ···················· | ···················· | ···················· |
| AB | 8296 | hCoV-19/USA/CO-CDPHE-2100010222/2020\|EPI_ISL_77129 | ···················· | ···················· | ···················· | ···················· | ···················· | ···················· |
| AB | 8297 | hCoV-19/USA/CO-CDPHE-2100010692/2020\|EPI_ISL_77129 | ···················· | ···················· | ···················· | ···················· | ···················· | ···················· |
| AB | 8298 | hCoV-19/USA/CO-CDPHE-2100012955/2020\|EPI_ISL_77129 | ···················· | ···················· | ···················· | ···················· | ···················· | ···················· |
| AB | 8299 | hCoV-19/USA/CO-CDPHE-2100012987/2020\|EPI_ISL_77129 | ········T··········· | ·A·················· | ···················· | ···················· | ···················· | ···················· |
| AB | 8300 | hCoV-19/USA/CO-CDPHE-2100013339/2020\|EPI_ISL_77129 | ···················· | ·A·················· | ···················· | ···················· | ···················· | ···················· |
| AB | 8301 | hCoV-19/Germany/HH-hpi-p1691/2020\|EPI_ISL_776100\|2 | ···················· | ···················· | ···················· | ···················· | ···················· | ···················· |
| AB | 8302 | hCoV-19/Germany/HH-hpi-p1696/2020\|EPI_ISL_776101\|2 | ···················· | ···················· | ···················· | ···················· | ···················· | ···················· |
| AB | 8303 | hCoV-19/Germany/HH-hpi-p1719/2020\|EPI_ISL_776102\|2 | ···················· | ···················· | ···················· | ···················· | ···················· | ···················· |
| AB | 8304 | hCoV-19/Germany/HH-hpi-p1720/2020\|EPI_ISL_776103\|2 | ···················· | ···················· | ···················· | ···················· | ···················· | ···················· |
| AB | 8305 | hCoV-19/Germany/HH-hpi-p1724/2020\|EPI_ISL_776104\|2 | ···················· | ···················· | ···················· | ···················· | ···················· | ···················· |
| AB | 8306 | hCoV-19/Germany/HH-hpi-p1725/2020\|EPI_ISL_776105\|2 | ···················· | ···················· | ···················· | ···················· | ···················· | ···················· |
| AB | 8307 | hCoV-19/Germany/HH-hpi-p1726/2020\|EPI_ISL_776106\|2 | ···················· | ···················· | ···················· | ···················· | ···················· | ···················· |
| AB | 8308 | hCoV-19/Germany/HH-hpi-p1739/2020\|EPI_ISL_776118\|2 | ···················· | ·A·················· | ···················· | ···················· | ···················· | ···················· |
| AB | 8309 | hCoV-19/Germany/HH-hpi-p1740/2020\|EPI_ISL_776119\|2 | ···················· | ···················· | ···················· | ···················· | ···················· | ···················· |
| AB | 8310 | hCoV-19/Germany/HH-hpi-p1742/2020\|EPI_ISL_776120\|2 | ···················· | ···················· | ···················· | ···················· | ···················· | ···················· |
| AB | 8311 | hCoV-19/Germany/HH-hpi-p1748/2020\|EPI_ISL_776125\|2 | ···················· | ···················· | ···················· | ···················· | ···················· | ···················· |
| AB | 8312 | hCoV-19/Germany/HH-hpi-p1751/2020\|EPI_ISL_776126\|2 | ···················· | ···················· | ···················· | ···················· | ···················· | ···················· |
| AB | 8313 | hCoV-19/Germany/HH-hpi-p1752/2020\|EPI_ISL_776127\|2 | ···················· | ···················· | ···················· | ···················· | ···················· | ···················· |
| AB | 8314 | hCoV-19/Germany/HH-hpi-p1753/2020\|EPI_ISL_776128\|2 | ···················· | ···················· | ···················· | ···················· | ···················· | ···················· |
| AB | 8315 | hCoV-19/Germany/HH-hpi-p1754/2020\|EPI_ISL_776129\|2 | ···················· | ···················· | ···················· | ···················· | ···················· | ···················· |
| AB | 8316 | hCoV-19/Germany/HH-hpi-p1755/2020\|EPI_ISL_776130\|2 | ···················· | ···················· | ···················· | ···················· | ···················· | ···················· |
| AB | 8317 | hCoV-19/Germany/HH-hpi-p1756/2020\|EPI_ISL_776131\|2 | ···················· | ···················· | ···················· | ···················· | ···················· | ···················· |
| AB | 8318 | hCoV-19/Germany/HH-hpi-p1757/2020\|EPI_ISL_776132\|2 | ···················· | ···················· | ···················· | ···················· | ···················· | ···················· |
| AB | 8319 | hCoV-19/Germany/HH-hpi-p1758/2020\|EPI_ISL_776133\|2 | ···················· | ···················· | ···················· | ···················· | ···················· | ···················· |
| AB | 8320 | hCoV-19/Germany/HH-hpi-p1759/2020\|EPI_ISL_776134\|2 | ···················· | ···················· | ···················· | ···················· | ···················· | ···················· |
| AB | 8321 | hCoV-19/Germany/HH-hpi-p1760/2020\|EPI_ISL_776135\|2 | ···················· | ···················· | ···················· | ···················· | ···················· | ···················· |
| AB | 8322 | hCoV-19/Germany/HH-hpi-p1762/2020\|EPI_ISL_776136\|2 | ···················· | ···················· | ···················· | ···················· | ···················· | ···················· |
| AB | 8323 | hCoV-19/Germany/HH-hpi-p1763/2020\|EPI_ISL_776137\|2 | ···················· | ···················· | ···················· | ···················· | ···················· | ···················· |
| AB | 8324 | hCoV-19/Germany/HH-hpi-p1764/2020\|EPI_ISL_776138\|2 | ···················· | ···················· | ···················· | ···················· | ···················· | ···················· |
| AB | 8325 | hCoV-19/Germany/HH-hpi-p1765/2020\|EPI_ISL_776139\|2 | ···················· | ···················· | ···················· | ···················· | ···················· | ···················· |
| AB | 8326 | hCoV-19/Germany/HH-hpi-p1766/2020\|EPI_ISL_776140\|2 | ···················· | ···················· | ···················· | ···················· | ···················· | ···················· |
| AB | 8327 | hCoV-19/Germany/HH-hpi-p1767/2020\|EPI_ISL_776141\|2 | ···················· | ···················· | ···················· | ···················· | ···················· | ···················· |
| AB | 8328 | hCoV-19/Germany/HH-hpi-p1768/2020\|EPI_ISL_776142\|2 | ···················· | ···················· | ···················· | ···················· | ···················· | ···················· |
| AB | 8329 | hCoV-19/Germany/HH-hpi-p1769/2020\|EPI_ISL_776143\|2 | ···················· | ···················· | ···················· | ···················· | ···················· | ···················· |
| AB | 8330 | hCoV-19/Germany/HH-hpi-p1770/2020\|EPI_ISL_776144\|2 | ···················· | ···················· | ···················· | ···················· | ···················· | ···················· |
| AB | 8331 | hCoV-19/Germany/HH-hpi-p1771/2020\|EPI_ISL_776145\|2 | ···················· | ···················· | ···················· | ···················· | ···················· | ···················· |
| AB | 8332 | hCoV-19/Germany/HH-hpi-p1772/2020\|EPI_ISL_776146\|2 | ···················· | ···················· | ···················· | ···················· | ···················· | ···················· |
| AB | 8333 | hCoV-19/Germany/HH-hpi-p1773/2020\|EPI_ISL_776147\|2 | ···················· | ···················· | ···················· | ···················· | ···················· | ···················· |
| AB | 8334 | hCoV-19/Germany/HH-hpi-p1775/2020\|EPI_ISL_776148\|2 | ···················· | ···················· | ···················· | ···················· | ···················· | ···················· |
| AB | 8335 | hCoV-19/Germany/HH-hpi-p1776/2020\|EPI_ISL_776149\|2 | ···················· | ···················· | ···················· | ···················· | ···················· | ···················· |
| AB | 8336 | hCoV-19/Germany/HH-hpi-p1777/2020\|EPI_ISL_776150\|2 | ···················· | ···················· | ···················· | ···················· | ···················· | ···················· |
| AB | 8337 | hCoV-19/Germany/HH-hpi-p1778/2020\|EPI_ISL_776151\|2 | ···················· | ···················· | ···················· | ···················· | ···················· | ···················· |
| AB | 8338 | hCoV-19/Germany/HH-hpi-p1779/2020\|EPI_ISL_776152\|2 | ···················· | ···················· | ···················· | ···················· | ···················· | ···················· |
| AB | 8339 | hCoV-19/Germany/HH-hpi-p1780/2020\|EPI_ISL_776153\|2 | ···················· | ·············A······ | ···················· | ···················· | ···················· | ···················· |
| AB | 8340 | hCoV-19/Germany/HH-hpi-p1783/2020\|EPI_ISL_776154\|2 | ···················· | ···················· | ···················· | ···················· | ···················· | ···················· |
| AB | 8341 | hCoV-19/Germany/HH-hpi-p1784/2020\|EPI_ISL_776155\|2 | ···················· | ···················· | ···················· | ···················· | ···················· | ···················· |
| AB | 8342 | hCoV-19/Germany/HH-hpi-p1785/2020\|EPI_ISL_776156\|2 | ···················· | ···················· | ···················· | ···················· | ···················· | ···················· |
| AB | 8343 | hCoV-19/Germany/HH-hpi-p1786/2020\|EPI_ISL_776157\|2 | ···················· | ···················· | ···················· | ···················· | ···················· | ···················· |
| AB | 8344 | hCoV-19/Germany/HH-hpi-p1787/2020\|EPI_ISL_776158\|2 | ···················· | ···················· | ···················· | ···················· | ···················· | ···················· |
| AB | 8345 | hCoV-19/Germany/HH-hpi-p1788/2020\|EPI_ISL_776159\|2 | ···················· | ···················· | ···················· | ···················· | ···················· | ···················· |
| AB | 8346 | hCoV-19/Germany/HH-hpi-p1789/2020\|EPI_ISL_776160\|2 | ···················· | ···················· | ···················· | ···················· | ···················· | ···················· |
| AB | 8347 | hCoV-19/Germany/HH-hpi-p1790/2020\|EPI_ISL_776161\|2 | ···················· | ···················· | ···················· | ···················· | ···················· | ···················· |
| AB | 8348 | hCoV-19/Germany/HH-hpi-p1791/2020\|EPI_ISL_776162\|2 | ···················· | ···················· | ···················· | ···················· | ···················· | ···················· |
| AB | 8349 | hCoV-19/Germany/HH-hpi-p1793/2020\|EPI_ISL_776163\|2 | ···················· | ···················· | ···················· | ···················· | ···················· | ···················· |
| AB | 8350 | hCoV-19/Germany/HH-hpi-p1795/2020\|EPI_ISL_776164\|2 | ···················· | ···················· | ···················· | ···················· | ···················· | ···················· |
| AB | 8352 | hCoV-19/Germany/HH-hpi-p1797/2020\|EPI_ISL_776166\|2 | ···················· | ···················· | ···················· | ···················· | ···················· | ···················· |
| AB | 8353 | hCoV-19/Germany/HH-hpi-p1799/2020\|EPI_ISL_776167\|2 | ···················· | ···················· | ···················· | ···················· | ···················· | ···················· |
| AB | 8354 | hCoV-19/Germany/HH-hpi-p1803/2020\|EPI_ISL_776168\|2 | ···················· | ···················· | ···················· | ···················· | ···················· | ···················· |
| AB | 8355 | hCoV-19/Germany/HH-hpi-p1810/2020\|EPI_ISL_776169\|2 | ···················· | ···················· | ···················· | ···················· | ···················· | ···················· |
| AB | 8356 | hCoV-19/England/CAMC-C997A1/2020\|EPI_ISL_777409\|20 | ···················· | ···················· | ···················· | ···················· | ···················· | ···················· |
| AB | 8357 | hCoV-19/England/QEUH-C9475B/2020\|EPI_ISL_777417\|20 | ···················· | ···················· | ···················· | ···················· | ······T············· | ···················· |
| AB | 8358 | hCoV-19/England/CAMC-C988FA/2020\|EPI_ISL_777418\|20 | ···················· | ···················· | ···················· | ···················· | ···················· | ···················· |
| AB | 8359 | hCoV-19/England/ALDP-C8E569/2020\|EPI_ISL_777424\|20 | ···················· | ···················· | ···················· | ···················· | ···················· | ···················· |
| AB | 8360 | hCoV-19/England/CAMC-C98E43/2020\|EPI_ISL_777428\|20 | ···················· | ···················· | ···················· | ···················· | ···················· | ···················· |
| AB | 8361 | hCoV-19/England/ALDP-C8DEBA/2020\|EPI_ISL_777430\|20 | ···················· | ···················· | ···················· | ···················· | ···················· | ···················· |
| AB | 8362 | hCoV-19/England/CAMC-C990C4/2020\|EPI_ISL_777433\|20 | ···················· | ···················· | ···················· | ···················· | ···················· | ···················· |

| | | | | | | | | |
|---|---|---|---|---|---|---|---|---|
| AB | 8363 | hCoV-19/England/CAMC-C985C6/2020\|EPI_ISL_777434\|20 | ···················· | ···················· | ····················· | ···················· | ···················· | ···················· |
| AB | 8364 | hCoV-19/England/QEUH-C940E7/2020\|EPI_ISL_777436\|20 | ···················· | ···················· | ····················· | ···················· | ···················· | ···················· |
| AB | 8365 | hCoV-19/England/QEUH-C9480D/2020\|EPI_ISL_777459\|20 | ···················· | ···················· | ····················· | ···················· | ···················· | ···················· |
| AB | 8366 | hCoV-19/England/CAMC-C9899D/2020\|EPI_ISL_777460\|20 | ···················· | ·A·················· | ····················· | ···················· | ···················· | ···A················ |
| AB | 8367 | hCoV-19/England/QEUH-C961A6/2020\|EPI_ISL_777461\|20 | ···················· | ···················· | ····················· | ···················· | ······T············· | ···················· |
| AB | 8368 | hCoV-19/England/ALDP-C8D834/2020\|EPI_ISL_777470\|20 | ···················· | ···················· | ····················· | ···················· | ···················· | ···················· |
| AB | 8369 | hCoV-19/England/QEUH-C93F18/2020\|EPI_ISL_777474\|20 | ···················· | ···················· | ····················· | ···················· | ···················· | ···················· |
| AB | 8370 | hCoV-19/England/QEUH-C93F27/2020\|EPI_ISL_777475\|20 | ···················· | ···················· | ····················· | ···················· | ···················· | ···················· |
| AB | 8371 | hCoV-19/England/CAMC-C98A9A/2020\|EPI_ISL_777478\|20 | ···················· | ···················· | ····················· | ···················· | ···················· | ···················· |
| AB | 8372 | hCoV-19/England/CAMC-C98B5B/2020\|EPI_ISL_777480\|20 | ···················· | ···················· | ····················· | ···················· | ···················· | ···················· |
| AB | 8373 | hCoV-19/England/CAMC-C99440/2020\|EPI_ISL_777482\|20 | ···················· | ·A·················· | ····················· | ···················· | ···················· | ···················· |
| AB | 8374 | hCoV-19/England/QEUH-C9615B/2020\|EPI_ISL_777484\|20 | ···················· | ···················· | ····················· | ···················· | ···················· | ···················· |
| AB | 8375 | hCoV-19/England/CAMC-C99361/2020\|EPI_ISL_777488\|20 | ···················· | ···················· | ····················· | ···················· | ···················· | ···················· |
| AB | 8377 | hCoV-19/England/QEUH-C93F63/2020\|EPI_ISL_777493\|20 | ···················· | ···················· | ····················· | ···················· | ···················· | ···················· |
| AB | 8378 | hCoV-19/England/QEUH-C93B6F/2020\|EPI_ISL_777497\|20 | ···················· | ···················· | ····················· | ···················· | ···················· | ···················· |
| AB | 8379 | hCoV-19/Italy/CAM-TIGEM-570/2020\|EPI_ISL_778700\|20 | ···················· | ···················· | ····················· | ···················· | ···················· | ···················· |
| AB | 8380 | hCoV-19/Italy/CAM-TIGEM-571/2020\|EPI_ISL_778701\|20 | ···················· | ···················· | ····················· | ···················· | ···················· | ···················· |
| AB | 8381 | hCoV-19/Italy/CAM-TIGEM-572/2020\|EPI_ISL_778702\|20 | ···················· | ···················· | ····················· | ···················· | ···················· | ···················· |
| AB | 8382 | hCoV-19/Italy/CAM-TIGEM-388/2020\|EPI_ISL_778704\|20 | ···················· | ···················· | ····················· | ···················· | ······T············· | ···················· |
| AB | 8383 | hCoV-19/Italy/CAM-TIGEM-394/2020\|EPI_ISL_778706\|20 | ···················· | ···················· | ····················· | ···················· | ···················· | ···················· |
| AB | 8384 | hCoV-19/Italy/CAM-TIGEM-400/2020\|EPI_ISL_778707\|20 | ···················· | ···················· | ····················· | ···················· | ······T············· | ···················· |
| AB | 8385 | hCoV-19/Italy/CAM-TIGEM-412/2020\|EPI_ISL_778708\|20 | ···················· | ···················· | ····················· | ···················· | ······T············· | ···················· |
| AB | 8386 | hCoV-19/Italy/CAM-TIGEM-411/2020\|EPI_ISL_778709\|20 | ···················· | ···················· | ····················· | ···················· | ······T············· | ···················· |
| AB | 8387 | hCoV-19/Italy/CAM-TIGEM-391/2020\|EPI_ISL_778710\|20 | ···················· | ···················· | ····················· | ···················· | ······T············· | ···················· |
| AB | 8388 | hCoV-19/Italy/CAM-TIGEM-414/2020\|EPI_ISL_778711\|20 | ···················· | ···················· | ····················· | ···················· | ······T············· | ···················· |
| AB | 8389 | hCoV-19/Italy/CAM-TIGEM-392/2020\|EPI_ISL_778713\|20 | ···················· | ···················· | ····················· | ···················· | ······T············· | ···················· |
| AB | 8390 | hCoV-19/Italy/CAM-TIGEM-420/2020\|EPI_ISL_778720\|20 | ···················· | ···················· | ····················· | ···················· | ······T············· | ···················· |
| AB | 8391 | hCoV-19/Italy/CAM-TIGEM-413/2020\|EPI_ISL_778721\|20 | ···················· | ···················· | ····················· | ···················· | ···················· | ···················· |
| AB | 8392 | hCoV-19/Italy/CAM-TIGEM-561/2020\|EPI_ISL_778722\|20 | ···················· | ···················· | ····················· | ···················· | ···················· | ···················· |
| AB | 8393 | hCoV-19/Italy/CAM-TIGEM-557/2020\|EPI_ISL_778725\|20 | ···················· | ···················· | ····················· | ···················· | ···················· | ···················· |
| AB | 8394 | hCoV-19/Italy/CAM-TIGEM-554/2020\|EPI_ISL_778726\|20 | ···················· | ···················· | ····················· | ···················· | ···················· | ···················· |
| AB | 8395 | hCoV-19/Italy/CAM-TIGEM-574/2020\|EPI_ISL_778727\|20 | ···················· | ···················· | ····················· | ···················· | ···················· | ···················· |
| AB | 8396 | hCoV-19/Italy/CAM-TIGEM-575/2020\|EPI_ISL_778728\|20 | ···················· | ···················· | ····················· | ···················· | ···················· | ···················· |
| AB | 8397 | hCoV-19/Italy/CAM-TIGEM-545/2020\|EPI_ISL_778729\|20 | ···················· | ···················· | ····················· | ···················· | ···················· | ···················· |
| AB | 8398 | hCoV-19/Italy/CAM-TIGEM-552/2020\|EPI_ISL_778730\|20 | ···················· | ···················· | ····················· | ···················· | ···················· | ···················· |
| AB | 8399 | hCoV-19/Italy/CAM-TIGEM-551/2020\|EPI_ISL_778731\|20 | ···················· | ···················· | ····················· | ···················· | ···················· | ···················· |
| AB | 8400 | hCoV-19/Italy/CAM-TIGEM-550/2020\|EPI_ISL_778732\|20 | ···················· | ···················· | ····················· | ···················· | ···················· | ···················· |
| AB | 8401 | hCoV-19/Italy/CAM-TIGEM-555/2020\|EPI_ISL_778733\|20 | ···················· | ···················· | ····················· | ···················· | ···················· | ···················· |
| AB | 8402 | hCoV-19/Italy/CAM-TIGEM-547/2020\|EPI_ISL_778734\|20 | ···················· | ···················· | ····················· | ···················· | ···················· | ···················· |
| AB | 8403 | hCoV-19/Italy/CAM-TIGEM-558/2020\|EPI_ISL_778735\|20 | ···················· | ···················· | ····················· | ···················· | ···················· | ···················· |
| AB | 8404 | hCoV-19/Italy/CAM-TIGEM-562/2020\|EPI_ISL_778736\|20 | ···················· | ···················· | ····················· | ···················· | ···················· | ···················· |
| AB | 8405 | hCoV-19/Italy/CAM-TIGEM-421/2020\|EPI_ISL_778737\|20 | ···················· | ···················· | ····················· | ···················· | ···················· | ···················· |
| AB | 8406 | hCoV-19/Italy/CAM-TIGEM-430/2020\|EPI_ISL_778738\|20 | ···················· | ···················· | ····················· | ···················· | ···················· | ···················· |
| AB | 8407 | hCoV-19/Italy/CAM-TIGEM-524/2020\|EPI_ISL_778739\|20 | ···················· | ···················· | ····················· | ···················· | ···················· | ···················· |
| AB | 8408 | hCoV-19/Italy/CAM-TIGEM-526/2020\|EPI_ISL_778740\|20 | ···················· | ···················· | ····················· | ···················· | ···················· | ···················· |
| AB | 8409 | hCoV-19/Italy/CAM-TIGEM-527/2020\|EPI_ISL_778741\|20 | ···················· | ···················· | ····················· | ···················· | ···················· | ···················· |
| AB | 8410 | hCoV-19/Italy/CAM-TIGEM-530/2020\|EPI_ISL_778742\|20 | ···················· | ···················· | ····················· | ···················· | ···················· | ···················· |
| AB | 8411 | hCoV-19/Italy/CAM-TIGEM-531/2020\|EPI_ISL_778743\|20 | ···················· | ···················· | ····················· | ···················· | ···················· | ···················· |
| AB | 8412 | hCoV-19/Italy/CAM-TIGEM-532/2020\|EPI_ISL_778744\|20 | ···················· | ···················· | ····················· | ···················· | ···················· | ···················· |
| AB | 8413 | hCoV-19/Italy/CAM-TIGEM-533/2020\|EPI_ISL_778745\|20 | ···················· | ···················· | ····················· | ···················· | ···················· | ···················· |
| AB | 8414 | hCoV-19/Italy/CAM-TIGEM-534/2020\|EPI_ISL_778746\|20 | ···················· | ···················· | ····················· | ···················· | ···················· | ···················· |
| AB | 8415 | hCoV-19/Italy/CAM-TIGEM-431/2020\|EPI_ISL_778747\|20 | ···················· | ···················· | ····················· | ···················· | ···················· | ···················· |
| AB | 8416 | hCoV-19/Italy/CAM-TIGEM-537/2020\|EPI_ISL_778749\|20 | ···················· | ···················· | ····················· | ···················· | ···················· | ···················· |
| AB | 8417 | hCoV-19/Italy/CAM-TIGEM-540/2020\|EPI_ISL_778750\|20 | ···················· | ···················· | ····················· | ···················· | ···················· | ···················· |
| AB | 8418 | hCoV-19/Italy/CAM-TIGEM-541/2020\|EPI_ISL_778751\|20 | ···················· | ···················· | ····················· | ···················· | ···················· | ···················· |
| AB | 8419 | hCoV-19/Italy/CAM-TIGEM-434/2020\|EPI_ISL_778752\|20 | ···················· | ···················· | ····················· | ···················· | ···················· | ···················· |
| AB | 8420 | hCoV-19/Italy/CAM-TIGEM-544/2020\|EPI_ISL_778753\|20 | ···················· | ···················· | ····················· | ···················· | ···················· | ···················· |
| AB | 8421 | hCoV-19/Italy/CAM-TIGEM-435/2020\|EPI_ISL_778754\|20 | ···················· | ···················· | ····················· | ···················· | ···················· | ···················· |
| AB | 8422 | hCoV-19/Italy/CAM-TIGEM-437/2020\|EPI_ISL_778756\|20 | ···················· | ···················· | ····················· | ···················· | ···················· | ···················· |
| AB | 8423 | hCoV-19/Italy/CAM-TIGEM-439/2020\|EPI_ISL_778758\|20 | ···················· | ···················· | ····················· | ···················· | ···················· | ···················· |
| AB | 8424 | hCoV-19/Italy/CAM-TIGEM-440/2020\|EPI_ISL_778759\|20 | ···················· | ···················· | ····················· | ···················· | ···················· | ···················· |
| AB | 8425 | hCoV-19/Italy/CAM-TIGEM-441/2020\|EPI_ISL_778760\|20 | ···················· | ···················· | ····················· | ···················· | ···················· | ···················· |
| AB | 8426 | hCoV-19/Italy/CAM-TIGEM-422/2020\|EPI_ISL_778761\|20 | ···················· | ···················· | ····················· | ···················· | ···················· | ···················· |
| AB | 8427 | hCoV-19/Italy/CAM-TIGEM-442/2020\|EPI_ISL_778762\|20 | ···················· | ···················· | ····················· | ···················· | ···················· | ···················· |
| AB | 8428 | hCoV-19/Italy/CAM-TIGEM-444/2020\|EPI_ISL_778763\|20 | ···················· | ···················· | ····················· | ···················· | ···················· | ···················· |
| AB | 8429 | hCoV-19/Italy/CAM-TIGEM-445/2020\|EPI_ISL_778764\|20 | ···················· | ···················· | ····················· | A··················· | ···················· | ···················· |
| AB | 8430 | hCoV-19/Italy/CAM-TIGEM-446/2020\|EPI_ISL_778765\|20 | ···················· | ···················· | ····················· | ···················· | ···················· | ···················· |
| AB | 8431 | hCoV-19/Italy/CAM-TIGEM-447/2020\|EPI_ISL_778766\|20 | ···················· | ···················· | ····················· | ···················· | ···················· | ···················· |
| AB | 8432 | hCoV-19/Italy/CAM-TIGEM-448/2020\|EPI_ISL_778767\|20 | ···················· | ···················· | ····················· | ···················· | ···················· | ···················· |
| AB | 8433 | hCoV-19/Italy/CAM-TIGEM-449/2020\|EPI_ISL_778768\|20 | ···················· | ···················· | ····················· | ···················· | ···················· | ···················· |
| AB | 8434 | hCoV-19/Italy/CAM-TIGEM-473/2020\|EPI_ISL_778780\|20 | ···················· | ···················· | ····················· | A··················· | ···················· | ···················· |
| AB | 8435 | hCoV-19/Italy/CAM-TIGEM-474/2020\|EPI_ISL_778781\|20 | ···················· | ···················· | ····················· | ···················· | ···················· | ···················· |
| AB | 8436 | hCoV-19/Italy/CAM-TIGEM-478/2020\|EPI_ISL_778782\|20 | ···················· | ···················· | ····················· | ···················· | ···················· | ···················· |
| AB | 8437 | hCoV-19/Italy/CAM-TIGEM-484/2020\|EPI_ISL_778787\|20 | ···················· | ···················· | ····················· | ···················· | ···················· | ···················· |
| AB | 8438 | hCoV-19/Italy/CAM-TIGEM-486/2020\|EPI_ISL_778789\|20 | ···················· | ···················· | ····················· | ···················· | ···················· | ···················· |
| AB | 8439 | hCoV-19/Italy/CAM-TIGEM-487/2020\|EPI_ISL_778790\|20 | ···················· | ···················· | ····················· | ···················· | ···················· | ···················· |
| AB | 8440 | hCoV-19/Italy/CAM-TIGEM-488/2020\|EPI_ISL_778791\|20 | ···················· | ···················· | ····················· | ···················· | ···················· | ···················· |
| AB | 8441 | hCoV-19/Italy/CAM-TIGEM-489/2020\|EPI_ISL_778792\|20 | ···················· | ···················· | ····················· | ···················· | ···················· | ···················· |
| AB | 8442 | hCoV-19/Italy/CAM-TIGEM-490/2020\|EPI_ISL_778793\|20 | ···················· | ···················· | ····················· | ···················· | ···················· | ···················· |
| AB | 8443 | hCoV-19/Italy/CAM-TIGEM-494/2020\|EPI_ISL_778794\|20 | ···················· | ···················· | ····················· | ···················· | ···················· | ···················· |

AB 8444 hCoV-19/Italy/CAM-TIGEM-427/2020|EPI_ISL_778795|20 ···················· ···················· ····················· ···················· ···················· ····················
AB 8445 hCoV-19/Italy/CAM-TIGEM-495/2020|EPI_ISL_778796|20 ···················· ···················· ····················· ···················· ···················· ····················
AB 8446 hCoV-19/Italy/CAM-TIGEM-497/2020|EPI_ISL_778797|20 ···················· ···················· ····················· ···················· ···················· ····················
AB 8447 hCoV-19/Italy/CAM-TIGEM-498/2020|EPI_ISL_778798|20 ···················· ···················· ····················· ···················· ···················· ····················
AB 8448 hCoV-19/Italy/CAM-TIGEM-499/2020|EPI_ISL_778799|20 ···················· ···················· ····················· ···················· ···················· ····················
AB 8449 hCoV-19/England/ALDP-CAE17E/2020|EPI_ISL_760600|20 ···················· ···················· ····················· ···················· ···················· ····················
AB 8450 hCoV-19/England/QEUH-CC236F/2020|EPI_ISL_760602|20 ···················· ···················· ····················· ···················· ···················· ····················
AB 8451 hCoV-19/England/QEUH-CC3568/2020|EPI_ISL_760604|20 ···················· ···················· ····················· ···················· ···················· ····················
AB 8452 hCoV-19/England/QEUH-CC2AB5/2020|EPI_ISL_760607|20 ···················· ···················· ····················· ···················· ···················· ····················
AB 8453 hCoV-19/England/ALDP-CBE7B5/2020|EPI_ISL_760609|20 ···················· ···················· ····················· ···················· ···················· ····················
AB 8454 hCoV-19/England/QEUH-CC47E9/2020|EPI_ISL_760616|20 ···················· ···················· ····················· ···················· ···················· ····················
AB 8455 hCoV-19/England/QEUH-CC4804/2020|EPI_ISL_760618|20 ···················· ···················· ····················· ···················· ···················· ····················
AB 8456 hCoV-19/England/QEUH-CC33C8/2020|EPI_ISL_760621|20 ···················· ···················· ····················· ···················· ···················· ····················
AB 8457 hCoV-19/England/ALDP-CBDE0C/2020|EPI_ISL_760623|20 ···················· ···················· ····················· ···················· ···················· ····················
AB 8458 hCoV-19/England/ALDP-CBE80D/2020|EPI_ISL_760625|20 ···················· ···················· ····················· ···················· ···················· ····················
AB 8459 hCoV-19/England/QEUH-CC2AA6/2020|EPI_ISL_760626|20 ···················· ···················· ····················· ···················· ···················· ····················
AB 8460 hCoV-19/England/ALDP-CBF59C/2020|EPI_ISL_760628|20 ···················· ···················· ····················· ···················· ···················· ····················
AB 8461 hCoV-19/England/QEUH-CC239C/2020|EPI_ISL_760629|20 ···················· ···················· ····················· ···················· ···················· ····················
AB 8462 hCoV-19/England/QEUH-CC21A1/2020|EPI_ISL_760632|20 ···················· ···················· ····················· ···················· ···················· ····················
AB 8463 hCoV-19/England/ALDP-CC046E/2020|EPI_ISL_760633|20 ···················· ···················· ····················· ···················· ···················· ····················
AB 8464 hCoV-19/England/QEUH-CC37F9/2020|EPI_ISL_760634|20 ···················· ···················· ····················· ···················· ···················· ····················
AB 8465 hCoV-19/England/QEUH-CC32E9/2020|EPI_ISL_760635|20 ···················· ···················· ····················· ···················· ···················· ····················
AB 8466 hCoV-19/England/QEUH-CC339B/2020|EPI_ISL_760636|20 ···················· ···················· ····················· ···················· ···················· ····················
AB 8467 hCoV-19/England/ALDP-CC05F2/2020|EPI_ISL_760637|20 ···················· ···················· ····················· ···················· ···················· ····················
AB 8468 hCoV-19/England/QEUH-CC3DAC/2020|EPI_ISL_760641|20 ···················· ···················· ····················· ···················· ···················· ····················
AB 8469 hCoV-19/England/QEUH-CC2E5F/2020|EPI_ISL_760643|20 ···················· ···················· ····················· ···················· ···················· ····················
AB 8470 hCoV-19/England/ALDP-CBE436/2020|EPI_ISL_760644|20 ···················· ···················· ····················· ···················· ···················· ····················
AB 8471 hCoV-19/England/QEUH-CC3322/2020|EPI_ISL_760645|20 ···················· ···················· ····················· ···················· ···················· ····················
AB 8472 hCoV-19/England/QEUH-CC2E8C/2020|EPI_ISL_760646|20 ···················· ···················· ····················· ···················· ···A················ ····················
AB 8473 hCoV-19/England/QEUH-CC29F4/2020|EPI_ISL_760648|20 ···················· ···················· ····················· ···················· ···················· ····················
AB 8474 hCoV-19/England/ALDP-CBF5C9/2020|EPI_ISL_760652|20 ···················· ···················· ····················· ···················· ···················· ····················
AB 8475 hCoV-19/England/QEUH-CC387E/2020|EPI_ISL_760655|20 ···················· ···················· ····················· ···················· ···················· ····················
AB 8476 hCoV-19/England/ALDP-CBE612/2020|EPI_ISL_760660|20 ···················· ···················· ····················· ···················· ···················· ····················
AB 8477 hCoV-19/England/QEUH-CC3DD9/2020|EPI_ISL_760662|20 ···················· ···················· ····················· ···················· ···················· ····················
AB 8478 hCoV-19/England/ALDP-CBE4DC/2020|EPI_ISL_760663|20 ···················· ···················· ····················· ···················· ···················· ····················
AB 8479 hCoV-19/England/ALDP-CBE83A/2020|EPI_ISL_760664|20 ···················· ···················· ····················· ···················· ···················· ····················
AB 8480 hCoV-19/England/ALDP-CBF3DE/2020|EPI_ISL_760670|20 ···················· ···················· ····················· ···················· ···················· ····················
AB 8481 hCoV-19/England/QEUH-CC351D/2020|EPI_ISL_760671|20 ···················· ·········T·········· ····················· ···················· ···················· ····················
AB 8482 hCoV-19/England/QEUH-CC33AA/2020|EPI_ISL_760674|20 ···················· ·A·················· ····················· ···················· ···················· ···A················
AB 8483 hCoV-19/England/QEUH-CC2A00/2020|EPI_ISL_760675|20 ···················· ···················· ····················· ···················· ···················· ····················
AB 8484 hCoV-19/England/QEUH-CC2253/2020|EPI_ISL_760676|20 ···················· ···················· ····················· ···················· ···················· ····················
AB 8485 hCoV-19/England/ALDP-CC060E/2020|EPI_ISL_760677|20 ···················· ···················· ····················· ···················· ···················· ····················
AB 8486 hCoV-19/England/ALDP-CBE551/2020|EPI_ISL_760680|20 ···················· ···················· ····················· ···················· ···················· ····················
AB 8487 hCoV-19/England/QEUH-CC4840/2020|EPI_ISL_760681|20 ···················· ···················· ····················· ···················· ···················· ····················
AB 8488 hCoV-19/England/QEUH-CC21FC/2020|EPI_ISL_760682|20 ···················· ···················· ····················· ···················· ···················· ····················
AB 8489 hCoV-19/England/QEUH-CC3823/2020|EPI_ISL_760683|20 ···················· ···················· ····················· ···················· ···················· ····················
AB 8490 hCoV-19/England/ALDP-CBE22D/2020|EPI_ISL_760693|20 ···················· ···················· ····················· ···················· ···················· ····················
AB 8491 hCoV-19/England/QEUH-CC2A4C/2020|EPI_ISL_760694|20 ···················· ·A·················· ····················· ···················· ···················· ···A················
AB 8492 hCoV-19/England/ALDP-CB6629/2020|EPI_ISL_761911|20 ···················· ···················· ····················· ···················· ···················· ····················
AB 8493 hCoV-19/England/ALDP-CB69E4/2020|EPI_ISL_761912|20 ···················· ···················· ····················· ···················· ······T············· ····················
AB 8494 hCoV-19/England/ALDP-CB3A20/2020|EPI_ISL_761914|20 ···················· ···················· ····················· ···················· ···················· ····················
AB 8495 hCoV-19/England/ALDP-CB5CDD/2020|EPI_ISL_761915|20 ···················· ···················· ····················· ···················· ···················· ····················
AB 8496 hCoV-19/England/ALDP-CB5CA0/2020|EPI_ISL_761916|20 ···················· ···················· ····················· ···················· ···················· ····················
AB 8497 hCoV-19/England/ALDP-CB5411/2020|EPI_ISL_761917|20 ···················· ···················· ····················· ···················· ···················· ····················
AB 8498 hCoV-19/England/ALDP-CB3756/2020|EPI_ISL_761919|20 ···················· ···················· ····················· ···················· ···················· ····················
AB 8499 hCoV-19/England/CAMC-CA0A08/2020|EPI_ISL_761922|20 ···················· ···················· ····················· ···················· ······T············· ····················
AB 8500 hCoV-19/England/CAMC-C99695/2020|EPI_ISL_761927|20 ···················· ···················· ····················· ···················· ···················· ····················
AB 8501 hCoV-19/England/CAMC-C98FF5/2020|EPI_ISL_761929|20 ···················· ···················· ····················· ···················· ···················· ····················
AB 8502 hCoV-19/England/QEUH-C92D3D/2020|EPI_ISL_761938|20 ···················· ···················· ····················· ···················· ···················· ····················
AB 8503 hCoV-19/England/QEUH-C92F46/2020|EPI_ISL_761957|20 ···················· ···················· ····················· ···················· ···················· ····················
AB 8504 hCoV-19/England/QEUH-C92F64/2020|EPI_ISL_761958|20 ···················· ···················· ····················· ···················· ···················· ····················
AB 8505 hCoV-19/England/QEUH-C92C12/2020|EPI_ISL_761963|20 ···················· ···················· ····················· ···················· ···················· ····················
AB 8506 hCoV-19/England/QEUH-C93103/2020|EPI_ISL_761965|20 ···················· ···················· ····················· ···················· ···················· ····················
AB 8507 hCoV-19/England/CAMC-C98FB9/2020|EPI_ISL_761967|20 ···················· ···················· ····················· ···················· ···················· ····················
AB 8508 hCoV-19/England/QEUH-C930E8/2020|EPI_ISL_761969|20 ···················· ···················· ····················· ···················· ···················· ····················
AB 8509 hCoV-19/England/QEUH-C93464/2020|EPI_ISL_761974|20 ···················· ···················· ····················· ···················· ···················· ····················
AB 8510 hCoV-19/England/QEUH-C92B60/2020|EPI_ISL_761978|20 ···················· ···················· ····················· ···················· ···················· ····················
AB 8511 hCoV-19/England/CAMC-C99659/2020|EPI_ISL_761979|20 ···················· ···················· ····················· ···················· ···················· ····················
AB 8512 hCoV-19/England/QEUH-C93D69/2020|EPI_ISL_761980|20 ···················· ···················· ····················· ···················· ···················· ····················
AB 8513 hCoV-19/England/CAMC-C98C67/2020|EPI_ISL_761981|20 ···················· ···················· ····················· ···················· ···················· ····················
AB 8514 hCoV-19/England/QEUH-C93288/2020|EPI_ISL_761983|20 ···················· ···················· ····················· ···················· ······T············· ····················
AB 8515 hCoV-19/England/QEUH-C933B2/2020|EPI_ISL_761989|20 ···················· ···················· ····················· ···················· ···················· ····················
AB 8516 hCoV-19/England/QEUH-C9319A/2020|EPI_ISL_761996|20 ···················· ···················· ····················· ···················· ······T············· ····················
AB 8517 hCoV-19/England/QEUH-C934A0/2020|EPI_ISL_761998|20 ···················· ···················· ····················· ···················· ···················· ····················
AB 8518 hCoV-19/France/BFC-IPP10929/2020|EPI_ISL_765477|20 ···················· ···················· ····················· ···················· ···················· ····················
AB 8519 hCoV-19/France/BFC-IPP10931/2020|EPI_ISL_765478|20 ···················· ···················· ····················· ···················· ···················· ····················
AB 8520 hCoV-19/France/GES-IPP10968/2020|EPI_ISL_765479|20 ···················· ···················· ····················· ···················· ···················· ····················
AB 8521 hCoV-19/England/CAMC-C7387D/2020|EPI_ISL_769006|20 ···················· ···················· ····················· ···················· ···················· ····················
AB 8522 hCoV-19/England/CAMC-C754B3/2020|EPI_ISL_769012|20 ···················· ···················· ····················· ···················· ···················· ····················
AB 8523 hCoV-19/England/CAMC-C71C55/2020|EPI_ISL_769016|20 ···················· ···················· ····················· ···················· ···················· ····················

| | | | | | | | |
|---|---|---|---|---|---|---|---|
| AB | 8524 hCoV-19/England/CAMC-C72207/2020\|EPI_ISL_769018\|20 | ···················· | ·A·················· | ···················· | ···················· | ···················· | ···A················ |
| AB | 8525 hCoV-19/England/CAMC-C74609/2020\|EPI_ISL_769028\|20 | ···················· | ···················· | ···················· | ···················· | ···················· | ···················· |
| AB | 8526 hCoV-19/England/CAMC-C71E7D/2020\|EPI_ISL_769041\|20 | ···················· | ···················· | ···················· | ···················· | ···················· | ···················· |
| AB | 8527 hCoV-19/England/CAMC-C737DA/2020\|EPI_ISL_769042\|20 | ···················· | ···················· | ···················· | ···················· | ···················· | ···················· |
| AB | 8528 hCoV-19/England/CAMC-C7202B/2020\|EPI_ISL_769046\|20 | ···················· | ···················· | ···················· | ···················· | ···················· | ···················· |
| AB | 8529 hCoV-19/England/CAMC-C72216/2020\|EPI_ISL_769047\|20 | ···················· | ···················· | ···················· | ···················· | ···················· | ···················· |
| AB | 8530 hCoV-19/England/CAMC-C72146/2020\|EPI_ISL_769050\|20 | ···················· | ···················· | ···················· | ···················· | ···················· | ···················· |
| AB | 8531 hCoV-19/England/CAMC-C7203A/2020\|EPI_ISL_769052\|20 | ···················· | ·A·················· | ···················· | ···················· | ···················· | ···A················ |
| AB | 8532 hCoV-19/England/CAMC-C738C8/2020\|EPI_ISL_769053\|20 | ···················· | ···················· | ···················· | ···················· | ······T············· | ···················· |
| AB | 8533 hCoV-19/England/CAMC-C70CB0/2020\|EPI_ISL_769060\|20 | ···················· | ···················· | ···················· | ···················· | ···················· | ···················· |
| AB | 8534 hCoV-19/England/CAMC-C755A1/2020\|EPI_ISL_769065\|20 | ···················· | ···················· | ···················· | ···················· | ···················· | ···················· |
| AB | 8535 hCoV-19/England/CAMC-C71F89/2020\|EPI_ISL_769066\|20 | ···················· | ···················· | ···················· | ···················· | ······T············· | ···················· |
| AB | 8536 hCoV-19/England/CAMC-C74821/2020\|EPI_ISL_769067\|20 | ···················· | ···················· | ···················· | ···················· | ···················· | ···················· |
| AB | 8537 hCoV-19/England/CAMC-C71BD0/2020\|EPI_ISL_769071\|20 | ···················· | ···················· | ···················· | ···················· | ···················· | ···················· |
| AB | 8538 hCoV-19/England/CAMC-C71CDD/2020\|EPI_ISL_769074\|20 | ···················· | ···················· | ···················· | ···················· | ···················· | ···················· |
| AB | 8539 hCoV-19/Costa Rica/INC-0158/2020\|EPI_ISL_770000\|20 | ···················· | ···················· | ···················· | ···················· | ···················· | ···················· |
| AB | 8540 hCoV-19/Costa Rica/INC-0160/2020\|EPI_ISL_770001\|20 | ···················· | ···················· | ···················· | ···················· | ···················· | ···················· |
| AB | 8541 hCoV-19/Costa Rica/INC-0150/2020\|EPI_ISL_770013\|20 | ···················· | ···················· | ···················· | ···················· | ···················· | ···················· |
| AB | 8542 hCoV-19/Costa Rica/INC-0151/2020\|EPI_ISL_770014\|20 | ···················· | ···················· | ···················· | ···················· | ···················· | ···················· |
| AB | 8543 hCoV-19/Costa Rica/INC-0164/2020\|EPI_ISL_770015\|20 | ···················· | ···················· | ···················· | ···················· | ···················· | ···················· |
| AB | 8544 hCoV-19/Costa Rica/INC-0166/2020\|EPI_ISL_770016\|20 | ···················· | ···················· | ···················· | ···················· | ···················· | ···················· |
| AB | 8545 hCoV-19/Costa Rica/INC-0167/2020\|EPI_ISL_770017\|20 | ···················· | ···················· | ···················· | ···················· | ···················· | ···················· |
| AB | 8546 hCoV-19/Costa Rica/INC-0169/2020\|EPI_ISL_770018\|20 | ···················· | ···················· | ···················· | ···················· | ···················· | ···················· |
| AB | 8547 hCoV-19/Costa Rica/INC-0170/2020\|EPI_ISL_770019\|20 | ···················· | ···················· | ···················· | ···················· | ···················· | ···················· |
| AB | 8548 hCoV-19/Costa Rica/INC-0179/2020\|EPI_ISL_770024\|20 | ···················· | ···················· | ···················· | ···················· | ···················· | ···················· |
| AB | 8549 hCoV-19/Costa Rica/INC-0165/2020\|EPI_ISL_770025\|20 | ···················· | ···················· | ···················· | ···················· | ···················· | ···················· |
| AB | 8550 hCoV-19/Costa Rica/INC-0180/2020\|EPI_ISL_770026\|20 | ···················· | ···················· | ···················· | ···················· | ···················· | ···················· |
| AB | 8551 hCoV-19/Costa Rica/INC-0156/2020\|EPI_ISL_770027\|20 | ···················· | ······A············· | ···················· | ···················· | ···················· | ···················· |
| AB | 8552 hCoV-19/Costa Rica/INC-0147/2020\|EPI_ISL_770028\|20 | ···················· | ···················· | ···················· | ···················· | ···················· | ···················· |
| AB | 8553 hCoV-19/Costa Rica/INC-0171/2020\|EPI_ISL_770029\|20 | ···················· | ···················· | ···················· | ···················· | ···················· | ···················· |
| AB | 8554 hCoV-19/Costa Rica/INC-0175/2020\|EPI_ISL_770030\|20 | ···················· | ···················· | ···················· | ···················· | ···················· | ···················· |
| AB | 8555 hCoV-19/Costa Rica/INC-0152/2020\|EPI_ISL_770031\|20 | ···················· | ···················· | ···················· | ···················· | ···················· | ···················· |
| AB | 8556 hCoV-19/Costa Rica/INC-0153/2020\|EPI_ISL_770032\|20 | ···················· | ···················· | ···················· | ···················· | ···················· | ···················· |
| AB | 8557 hCoV-19/Latvia/200/2020\|EPI_ISL_770033\|2020-09-21 | ···················· | ···················· | ···················· | ···················· | ···················· | ···················· |
| AB | 8558 hCoV-19/Latvia/201/2020\|EPI_ISL_770034\|2020-09-21 | ···················· | ···················· | ···················· | ···················· | ···················· | ···················· |
| AB | 8559 hCoV-19/Latvia/212/2020\|EPI_ISL_770035\|2020-09-16 | ···················· | ···················· | ···················· | ···················· | ···················· | ···················· |
| AB | 8560 hCoV-19/Latvia/198/2020\|EPI_ISL_770036\|2020-09-23 | ···················· | ···················· | ···················· | ···················· | ···················· | ···················· |
| AB | 8561 hCoV-19/Latvia/199/2020\|EPI_ISL_770037\|2020-09-23 | ···················· | ···················· | ···················· | ···················· | ···················· | ···················· |
| AB | 8562 hCoV-19/Latvia/202/2020\|EPI_ISL_770038\|2020-09-18 | ···················· | ···················· | ···················· | ···················· | ···················· | ···················· |
| AB | 8563 hCoV-19/Latvia/203/2020\|EPI_ISL_770039\|2020-09-10 | ···················· | ···················· | ···················· | ···················· | ···················· | ···················· |
| AB | 8564 hCoV-19/Latvia/204/2020\|EPI_ISL_770040\|2020-09-23 | ···················· | ···················· | ···················· | ···················· | ···················· | ···················· |
| AB | 8565 hCoV-19/Latvia/205/2020\|EPI_ISL_770041\|2020-09-21 | ···················· | ···················· | ···················· | ···················· | ···················· | ···················· |
| AB | 8566 hCoV-19/Latvia/206/2020\|EPI_ISL_770042\|2020-09-16 | ···················· | ···················· | ···················· | ···················· | ···················· | ···················· |
| AB | 8567 hCoV-19/Latvia/207/2020\|EPI_ISL_770043\|2020-09-14 | ···················· | ···················· | ···················· | ···················· | ···················· | ···················· |
| AB | 8568 hCoV-19/Latvia/208/2020\|EPI_ISL_770044\|2020-09-16 | ···················· | ···················· | ···················· | ···················· | ···················· | ···················· |
| AB | 8569 hCoV-19/Latvia/209/2020\|EPI_ISL_770045\|2020-09-19 | ···················· | ···················· | ···················· | ···················· | ···················· | ···················· |
| AB | 8570 hCoV-19/Latvia/210/2020\|EPI_ISL_770046\|2020-09-12 | ···················· | ·A·················· | ···················· | ···················· | ···················· | ···A················ |
| AB | 8571 hCoV-19/Latvia/211/2020\|EPI_ISL_770047\|2020-09-02 | ···················· | ···················· | ···················· | ···················· | ···················· | ···················· |
| AB | 8572 hCoV-19/Latvia/213/2020\|EPI_ISL_770048\|2020-09-15 | ···················· | ···················· | ···················· | ···················· | ···················· | ···················· |
| AB | 8573 hCoV-19/Latvia/214/2020\|EPI_ISL_770049\|2020-09-13 | ···················· | ···················· | ···················· | ···················· | ···················· | ···················· |
| AB | 8574 hCoV-19/Latvia/215/2020\|EPI_ISL_770050\|2020-09-03 | ···················· | ···················· | ···················· | ···················· | ···················· | ···················· |
| AB | 8575 hCoV-19/Latvia/216/2020\|EPI_ISL_770051\|2020-09-23 | ···················· | ···················· | ···················· | ···················· | ···················· | ···················· |
| AB | 8576 hCoV-19/Latvia/217/2020\|EPI_ISL_770052\|2020-09-15 | ···················· | ···················· | ···················· | ···················· | ···················· | ···················· |
| AB | 8577 hCoV-19/Latvia/218/2020\|EPI_ISL_770053\|2020-09-17 | ···················· | ···················· | ···················· | ···················· | ···················· | ···················· |
| AB | 8578 hCoV-19/Latvia/219/2020\|EPI_ISL_770054\|2020-09-16 | ···················· | ···················· | ···················· | ···················· | ···················· | ···················· |
| AB | 8579 hCoV-19/Latvia/220/2020\|EPI_ISL_770055\|2020-09-14 | ···················· | ···················· | ···················· | ···················· | ···················· | ···················· |
| AB | 8580 hCoV-19/Latvia/222/2020\|EPI_ISL_770056\|2020-09-15 | ···················· | ···················· | ···················· | ···················· | ···················· | ···················· |
| AB | 8581 hCoV-19/Latvia/223/2020\|EPI_ISL_770057\|2020-09-14 | ···················· | ···················· | ···················· | ···················· | ···················· | ···················· |
| AB | 8582 hCoV-19/Latvia/224/2020\|EPI_ISL_770058\|2020-09-10 | ···················· | ···················· | ···················· | ···················· | ···················· | ···················· |
| AB | 8583 hCoV-19/Latvia/225/2020\|EPI_ISL_770059\|2020-09-19 | ···················· | ···················· | ···················· | ···················· | ···················· | ···················· |
| AB | 8584 hCoV-19/Latvia/226/2020\|EPI_ISL_770060\|2020-09-13 | ···················· | ···················· | ···················· | ···················· | ···················· | ···················· |
| AB | 8585 hCoV-19/Latvia/227/2020\|EPI_ISL_770061\|2020-09-18 | ···················· | ···················· | ···················· | ···················· | ···················· | ···················· |
| AB | 8586 hCoV-19/Latvia/228/2020\|EPI_ISL_770062\|2020-09-22 | ···················· | ···················· | ···················· | ···················· | ···················· | ···················· |
| AB | 8587 hCoV-19/USA/WY-WPHL-20153006/2020\|EPI_ISL_770066\|2 | ···················· | ···················· | ···················· | ···················· | ···················· | ···················· |
| AB | 8588 hCoV-19/USA/WY-WPHL-20148300/2020\|EPI_ISL_770067\|2 | ···················· | ·A·················· | ···················· | ···················· | ···················· | ···················· |
| AB | 8589 hCoV-19/USA/CO-CDPHE-2100014199/2020\|EPI_ISL_77130 | ···················· | ···················· | ···················· | ···················· | ···················· | ···················· |
| AB | 8590 hCoV-19/USA/CO-CDPHE-2100014244/2020\|EPI_ISL_77130 | ···················· | ···················· | ···················· | ···················· | ···················· | ···················· |
| AB | 8591 hCoV-19/USA/CO-CDPHE-2100014301/2020\|EPI_ISL_77130 | ···················· | ···················· | ···················· | ···················· | ···················· | ···················· |
| AB | 8592 hCoV-19/USA/CO-CDPHE-2100014374/2020\|EPI_ISL_77130 | ···················· | ···················· | ···················· | ···················· | ···················· | ···················· |
| AB | 8593 hCoV-19/USA/CO-CDPHE-2100014440/2020\|EPI_ISL_77130 | ···················· | ···················· | ···················· | ···················· | ···················· | ···················· |
| AB | 8594 hCoV-19/USA/CO-CDPHE-2100014442/2020\|EPI_ISL_77130 | ···················· | ·A·················· | ···················· | ···················· | ···················· | ···················· |
| AB | 8595 hCoV-19/USA/CO-CDPHE-2100014448/2020\|EPI_ISL_77130 | ···················· | ···················· | ···················· | ···················· | ···················· | ···················· |
| AB | 8596 hCoV-19/USA/CO-CDPHE-2100014535/2020\|EPI_ISL_77130 | ···················· | ···················· | ···················· | ···················· | ···················· | ···················· |
| AB | 8597 hCoV-19/USA/CO-CDPHE-2100014869/2020\|EPI_ISL_77130 | ···················· | ···················· | ···················· | ···················· | ···················· | ···················· |
| AB | 8598 hCoV-19/USA/CO-CDPHE-2100015072/2020\|EPI_ISL_77130 | ···················· | ···················· | ···················· | ···················· | ···················· | ···················· |
| AB | 8599 hCoV-19/USA/CO-CDPHE-2100015096/2020\|EPI_ISL_77131 | ···················· | ···················· | ···················· | ···················· | ···················· | ···················· |
| AB | 8600 hCoV-19/USA/CO-CDPHE-2100015131/2020\|EPI_ISL_77131 | ···················· | ···················· | ···················· | ···················· | ···················· | ···················· |
| AB | 8601 hCoV-19/USA/CO-CDPHE-2100015182/2020\|EPI_ISL_77131 | ···················· | ···················· | ···················· | ···················· | ···················· | ···················· |
| AB | 8602 hCoV-19/USA/CO-CDPHE-2100015371/2020\|EPI_ISL_77131 | ···················· | ···················· | ···················· | ···················· | ···················· | ···················· |
| AB | 8603 hCoV-19/USA/CO-CDPHE-2100015712/2020\|EPI_ISL_77131 | ···················· | ···················· | ···················· | ···················· | ···················· | ···················· |

| | | | | | | | | |
|---|---|---|---|---|---|---|---|---|
| AB | 8604 hCoV-19/USA/CO-CDPHE-2100015805/2020\|EPI_ISL_77131 | ··················· | ·A················· | ··················· | ··················· | ··················· | ··················· | |
| AB | 8605 hCoV-19/USA/CO-CDPHE-2100015827/2020\|EPI_ISL_77131 | ··················· | ··················· | ··················· | ··················· | ··················· | ··················· | |
| AB | 8606 hCoV-19/USA/CO-CDPHE-2100015900/2020\|EPI_ISL_77131 | ··················· | ··················· | ··················· | ··················· | ··N················ | ··················· | |
| AB | 8607 hCoV-19/USA/CO-CDPHE-2100015910/2020\|EPI_ISL_77131 | ··················· | ·A················· | ··················· | ··················· | ··················· | ··················· | |
| AB | 8608 hCoV-19/USA/CO-CDPHE-2100015923/2020\|EPI_ISL_77131 | ··················· | ··················· | ··················· | ··················· | ··················· | ··················· | |
| AB | 8609 hCoV-19/USA/CO-CDPHE-2100015982/2020\|EPI_ISL_77132 | ··················· | ··················· | ··················· | ··················· | ··················· | ··················· | |
| AB | 8610 hCoV-19/USA/CO-CDPHE-2100015997/2020\|EPI_ISL_77132 | ··················· | ·A················· | ··················· | ··················· | ··················· | ··················· | |
| AB | 8611 hCoV-19/USA/CO-CDPHE-2100016166/2020\|EPI_ISL_77132 | ··················· | ··················· | ··················· | ··················· | ··················· | ··················· | |
| AB | 8613 hCoV-19/USA/CO-CDPHE-2100016535/2020\|EPI_ISL_77132 | ··················· | ··················· | ··················· | ··················· | ··················· | ··················· | |
| AB | 8614 hCoV-19/USA/CO-CDPHE-2100016669/2020\|EPI_ISL_77132 | ··················· | ·A················· | ··················· | ··················· | ··················· | ··················· | |
| AB | 8615 hCoV-19/USA/CO-CDPHE-2100016689/2020\|EPI_ISL_77132 | ··················· | ··················· | ··················· | ··················· | ··················· | ··················· | |
| AB | 8616 hCoV-19/USA/CO-CDPHE-2100021102/2020\|EPI_ISL_77132 | ··················· | ··················· | ··················· | ··················· | ··················· | ··················· | |
| AB | 8617 hCoV-19/USA/CO-CDPHE-2100021189/2020\|EPI_ISL_77132 | ··················· | ··················· | ··················· | ··················· | ··················· | ··················· | |
| AB | 8618 hCoV-19/USA/CO-CDPHE-2100022857/2020\|EPI_ISL_77132 | ··················· | ·A················· | ··················· | ··················· | ··················· | ··················· | |
| AB | 8619 hCoV-19/USA/CO-CDPHE-2100023264/2020\|EPI_ISL_77133 | ··················· | ··················· | ··················· | ··················· | ··················· | ··················· | |
| AB | 8620 hCoV-19/USA/CO-CDPHE-2100023395/2020\|EPI_ISL_77133 | ··················· | ··················· | ··················· | ··················· | ··················· | ··················· | |
| AB | 8621 hCoV-19/USA/CO-CDPHE-2100023473/2020\|EPI_ISL_77133 | ··················· | ··················· | ··················· | ··················· | ··················· | ··················· | |
| AB | 8622 hCoV-19/USA/CO-CDPHE-2100036501/2020\|EPI_ISL_77134 | ··················· | ··················· | ··················· | ··················· | ··················· | ··················· | |
| AB | 8626 hCoV-19/USA/CO-CDPHE-2100037907/2020\|EPI_ISL_77134 | ··················· | ··················· | ··················· | ··················· | NNN················ | ··················· | 1 |
| AB | 8627 hCoV-19/USA/CO-CDPHE-2100037996/2020\|EPI_ISL_77134 | ··················· | ··················· | ··················· | ··················· | ··················· | ··················· | |
| AB | 8628 hCoV-19/USA/CO-CDPHE-2100042687/2020\|EPI_ISL_77135 | ··················· | ··················· | ··················· | ··················· | ··················· | ··················· | |
| AB | 8630 hCoV-19/USA/CO-CDPHE-2100042730/2020\|EPI_ISL_77135 | ··················· | ··················· | ··················· | ··················· | ··················· | ··················· | |
| AB | 8631 hCoV-19/USA/CO-CDPHE-2100042764/2020\|EPI_ISL_77135 | ··················· | ··················· | ··················· | ··················· | ··················· | ··················· | |
| AB | 8632 hCoV-19/USA/CO-CDPHE-2100042785/2020\|EPI_ISL_77135 | ··················· | ··················· | ··················· | ··················· | ··················· | ··················· | |
| AB | 8633 hCoV-19/USA/CO-CDPHE-2100042884/2020\|EPI_ISL_77136 | ··················· | ·A················· | ··················· | ··················· | ··················· | ··················· | |
| AB | 8634 hCoV-19/USA/WA-S2917/2020\|EPI_ISL_771362\|2020-07-2 | ··················· | ··················· | ··················· | ··················· | ··················· | ··················· | |
| AB | 8635 hCoV-19/USA/WA-S2920/2020\|EPI_ISL_771365\|2020-07-2 | ··················· | ··················· | ··················· | ··················· | ··················· | ··················· | |
| AB | 8636 hCoV-19/Germany/HH-hpi-p1727/2020\|EPI_ISL_776107\|2 | ··················· | ··················· | ··················· | ··················· | ··················· | ··················· | |
| AB | 8637 hCoV-19/Germany/HH-hpi-p1728/2020\|EPI_ISL_776108\|2 | ··················· | ··················· | ··················· | ··················· | ··················· | ··················· | |
| AB | 8638 hCoV-19/Germany/HH-hpi-p1729/2020\|EPI_ISL_776109\|2 | ··················· | ··················· | ··················· | ··················· | ··················· | ··················· | |
| AB | 8639 hCoV-19/Germany/HH-hpi-p1730/2020\|EPI_ISL_776110\|2 | ··················· | ··················· | ··················· | ··················· | ··················· | ··················· | |
| AB | 8640 hCoV-19/Germany/HH-hpi-p1731/2020\|EPI_ISL_776111\|2 | ··················· | ··················· | ··················· | ··················· | ··················· | ··················· | |
| AB | 8641 hCoV-19/Germany/HH-hpi-p1732/2020\|EPI_ISL_776112\|2 | ··················· | ··················· | ··················· | ··················· | ··················· | ··················· | |
| AB | 8642 hCoV-19/Germany/HH-hpi-p1734/2020\|EPI_ISL_776113\|2 | ··················· | ··················· | ··················· | ··················· | ··················· | ··················· | |
| AB | 8643 hCoV-19/Germany/HH-hpi-p1735/2020\|EPI_ISL_776114\|2 | ··················· | ··················· | ··················· | ··················· | ··················· | ··················· | |
| AB | 8644 hCoV-19/Germany/HH-hpi-p1736/2020\|EPI_ISL_776115\|2 | ··················· | ··················· | ··················· | ··················· | ··················· | ··················· | |
| AB | 8645 hCoV-19/Germany/HH-hpi-p1737/2020\|EPI_ISL_776116\|2 | ··················· | ··················· | ··················· | ··················· | ··················· | ··················· | |
| AB | 8646 hCoV-19/Germany/HH-hpi-p1738/2020\|EPI_ISL_776117\|2 | ··················· | ··················· | ··················· | ··················· | ··················· | ··················· | |
| AB | 8647 hCoV-19/Germany/HH-hpi-p1743/2020\|EPI_ISL_776121\|2 | ··················· | ··················· | ··················· | ··················· | ··················· | ··················· | |
| AB | 8648 hCoV-19/Germany/HH-hpi-p1744/2020\|EPI_ISL_776122\|2 | ··················· | ··················· | ··················· | ··················· | ··················· | ··················· | |
| AB | 8649 hCoV-19/Germany/HH-hpi-p1745/2020\|EPI_ISL_776123\|2 | ··················· | ··················· | ··················· | ··················· | ··················· | ··················· | |
| AB | 8650 hCoV-19/Germany/HH-hpi-p1747/2020\|EPI_ISL_776124\|2 | ··················· | ··················· | ··················· | ··················· | ··················· | ··················· | |
| AB | 8651 hCoV-19/Germany/HH-hpi-p1811/2020\|EPI_ISL_776170\|2 | ··················· | ··················· | ··················· | ··················· | ··················· | ··················· | |
| AB | 8652 hCoV-19/Germany/HH-hpi-p1813/2020\|EPI_ISL_776171\|2 | ··················· | ··················· | ··················· | ··················· | ··················· | ··················· | |
| AB | 8653 hCoV-19/Germany/HH-hpi-p1814/2020\|EPI_ISL_776172\|2 | ··················· | ··················· | ··················· | ··················· | ··················· | ··················· | |
| AB | 8654 hCoV-19/Germany/HH-hpi-p1815/2020\|EPI_ISL_776173\|2 | ··················· | ··················· | ··················· | ··················· | ··················· | ··················· | |
| AB | 8655 hCoV-19/Germany/HH-hpi-p1818/2020\|EPI_ISL_776174\|2 | ··················· | ··················· | ··················· | ··················· | ··················· | ··················· | |
| AB | 8656 hCoV-19/Germany/HH-hpi-p1819/2020\|EPI_ISL_776175\|2 | ··················· | ··················· | ··················· | ··················· | ··················· | ··················· | |
| AB | 8657 hCoV-19/Germany/HH-hpi-p1820/2020\|EPI_ISL_776176\|2 | ··················· | ··················· | ··················· | ··················· | ··················· | ··················· | |
| AB | 8658 hCoV-19/Germany/HH-hpi-p1821/2020\|EPI_ISL_776177\|2 | ··················· | ··················· | ··················· | ··················· | ··················· | ··················· | |
| AB | 8659 hCoV-19/Germany/HH-hpi-p1822/2020\|EPI_ISL_776178\|2 | ··················· | ··················· | ··················· | ··················· | ··················· | ··················· | |
| AB | 8660 hCoV-19/Germany/HH-hpi-p1823/2020\|EPI_ISL_776179\|2 | ··················· | ··················· | ··················· | ··················· | ··················· | ··················· | |
| AB | 8661 hCoV-19/Germany/HH-hpi-p1824/2020\|EPI_ISL_776180\|2 | ··················· | ··················· | ··················· | ··················· | ··················· | ··················· | |
| AB | 8662 hCoV-19/Germany/HH-hpi-p1827/2020\|EPI_ISL_776181\|2 | ··················· | ··················· | ··················· | ··················· | ··················· | ··················· | |
| AB | 8663 hCoV-19/Germany/HH-hpi-p1828/2020\|EPI_ISL_776182\|2 | ··················· | ··················· | ··················· | ··················· | ··················· | ··················· | |
| AB | 8664 hCoV-19/Germany/HH-hpi-p1829/2020\|EPI_ISL_776183\|2 | ··················· | ··················· | ··················· | ··················· | ··················· | ··················· | |
| AB | 8665 hCoV-19/Germany/HH-hpi-p1830/2020\|EPI_ISL_776184\|2 | ··················· | ··················· | ··················· | ··················· | ··················· | ··················· | |
| AB | 8666 hCoV-19/Germany/HH-hpi-p1832/2020\|EPI_ISL_776185\|2 | ··················· | ··················· | ··················· | ··················· | ··················· | ··················· | |
| AB | 8667 hCoV-19/Germany/HH-hpi-p1833/2020\|EPI_ISL_776186\|2 | ··················· | ··················· | ··················· | ··················· | ··················· | ··················· | |
| AB | 8668 hCoV-19/Germany/HH-hpi-p1834/2020\|EPI_ISL_776187\|2 | ··················· | ··················· | ··················· | ··················· | ··················· | ··················· | |
| AB | 8669 hCoV-19/Germany/HH-hpi-p1835/2020\|EPI_ISL_776188\|2 | ··················· | ··················· | ··················· | ··················· | ··················· | ··················· | |
| AB | 8670 hCoV-19/Germany/HH-hpi-p1836/2020\|EPI_ISL_776189\|2 | ··················· | ··················· | ··················· | ··················· | ··················· | ··················· | |
| AB | 8671 hCoV-19/Germany/HH-hpi-p1837/2020\|EPI_ISL_776190\|2 | ··················· | ··················· | ··················· | ··················· | ··················· | ··················· | |
| AB | 8672 hCoV-19/Germany/HH-hpi-p1838/2020\|EPI_ISL_776191\|2 | ··················· | ··················· | ··················· | ··················· | ··················· | ··················· | |
| AB | 8673 hCoV-19/Germany/HH-hpi-p1839/2020\|EPI_ISL_776192\|2 | ··················· | ·A················· | ··················· | ··················· | ··················· | ··················· | |
| AB | 8674 hCoV-19/Germany/HH-hpi-p1842/2020\|EPI_ISL_776193\|2 | ··················· | ··················· | ··················· | ··················· | ··················· | ··················· | |
| AB | 8675 hCoV-19/Germany/HH-hpi-p1843/2020\|EPI_ISL_776194\|2 | ··················· | ··················· | ··················· | ··················· | ··················· | ··················· | |
| AB | 8676 hCoV-19/Germany/HH-hpi-p1844/2020\|EPI_ISL_776195\|2 | ··················· | ··················· | ··················· | ··················· | ··················· | ··················· | |
| AB | 8677 hCoV-19/Germany/HH-hpi-p1845/2020\|EPI_ISL_776196\|2 | ··················· | ··················· | ··················· | ··················· | ··················· | ··················· | |
| AB | 8678 hCoV-19/Germany/HH-hpi-p1846/2020\|EPI_ISL_776197\|2 | ··················· | ··················· | ··················· | ··················· | ··················· | ··················· | |
| AB | 8679 hCoV-19/Germany/HH-hpi-p1847/2020\|EPI_ISL_776198\|2 | ··················· | ··················· | ··················· | ··················· | ··················· | ··················· | |
| AB | 8680 hCoV-19/Germany/HH-hpi-p1848/2020\|EPI_ISL_776199\|2 | ··················· | ··················· | ··················· | ··················· | ··················· | ··················· | |
| AB | 8681 hCoV-19/England/QEUH-C947C4/2020\|EPI_ISL_777438\|20 | ··················· | ··················· | ··················· | ··················· | ··················· | ··················· | |
| AB | 8682 hCoV-19/England/CAMC-C9870C/2020\|EPI_ISL_777439\|20 | ··················· | ··················· | ··················· | ··················· | ··················· | ··················· | |
| AB | 8683 hCoV-19/England/QEUH-C93DF0/2020\|EPI_ISL_777444\|20 | ··················· | ··················· | ··················· | ··················· | ··················· | ··················· | |
| AB | 8684 hCoV-19/England/CAMC-C99404/2020\|EPI_ISL_777445\|20 | ··················· | ··················· | ··················· | ··················· | ··················· | ··················· | |
| AB | 8685 hCoV-19/England/QEUH-C93E57/2020\|EPI_ISL_777447\|20 | ··················· | ··················· | ··················· | ··················· | ··················· | ··················· | |
| AB | 8686 hCoV-19/England/ALDP-C8DA3E/2020\|EPI_ISL_777448\|20 | ··················· | ··················· | ··················· | ··················· | ··················· | ··················· | |
| AB | 8687 hCoV-19/England/CAMC-C99756/2020\|EPI_ISL_777453\|20 | ··················· | ··················· | ··················· | ··················· | ··················· | ··················· | |
| AB | 8688 hCoV-19/England/CAMC-C98AC7/2020\|EPI_ISL_777454\|20 | ··················· | ··················· | ··················· | ··················· | ·····T············· | ··················· | |

| | | | | | | | | | |
|---|---|---|---|---|---|---|---|---|---|
| AB | 8689 | hCoV-19/England/ALDP-C8DE6F/2020\|EPI_ISL_777455\|20 | ···················· | ···················· | ···················· | ···················· | ···················· | ···················· | |
| AB | 8690 | hCoV-19/England/ALDP-C8D98C/2020\|EPI_ISL_777458\|20 | ···················· | ·A·················· | ···················· | ···················· | ···················· | ···A················ | |
| AB | 8691 | hCoV-19/Italy/CAM-TIGEM-450/2020\|EPI_ISL_778769\|20 | ···················· | ···················· | ···················· | ···················· | ···················· | ···················· | |
| AB | 8692 | hCoV-19/Italy/CAM-TIGEM-423/2020\|EPI_ISL_778770\|20 | ···················· | ···················· | ···················· | ···················· | ···················· | ···················· | |
| AB | 8693 | hCoV-19/Italy/CAM-TIGEM-452/2020\|EPI_ISL_778771\|20 | ···················· | ···················· | ···················· | ···················· | ···················· | ···················· | |
| AB | 8694 | hCoV-19/Italy/CAM-TIGEM-454/2020\|EPI_ISL_778772\|20 | ···················· | ···················· | ···················· | ···················· | ···················· | ···················· | |
| AB | 8695 | hCoV-19/Italy/CAM-TIGEM-462/2020\|EPI_ISL_778774\|20 | ···················· | ···················· | ···················· | ···················· | ···················· | ···················· | |
| AB | 8696 | hCoV-19/Italy/CAM-TIGEM-466/2020\|EPI_ISL_778775\|20 | ···················· | ···················· | ···················· | ···················· | ···················· | ···················· | |
| AB | 8697 | hCoV-19/Italy/CAM-TIGEM-467/2020\|EPI_ISL_778776\|20 | ···················· | ···················· | ···················· | ···················· | ···················· | ···················· | |
| AB | 8698 | hCoV-19/Italy/CAM-TIGEM-471/2020\|EPI_ISL_778778\|20 | ···················· | ···················· | ···················· | ···················· | ···················· | ···················· | |
| AB | 8699 | hCoV-19/Italy/CAM-TIGEM-425/2020\|EPI_ISL_778779\|20 | ···················· | ···················· | ···················· | ···················· | ···················· | ···················· | |
| AB | 8700 | hCoV-19/Italy/CAM-TIGEM-479/2020\|EPI_ISL_778783\|20 | ···················· | ···················· | ···················· | ···················· | ···················· | ···················· | |
| AB | 8701 | hCoV-19/Italy/CAM-TIGEM-482/2020\|EPI_ISL_778784\|20 | ···················· | ···················· | ···················· | ···················· | ···················· | ···················· | |
| AB | 8702 | hCoV-19/Italy/CAM-TIGEM-483/2020\|EPI_ISL_778785\|20 | ···················· | ···················· | ···················· | ···················· | ···················· | ···················· | |
| AB | 8703 | hCoV-19/Italy/CAM-TIGEM-426/2020\|EPI_ISL_778786\|20 | ···················· | ···················· | ···················· | ···················· | ···················· | ···················· | |
| AB | 8704 | hCoV-19/Italy/CAM-TIGEM-485/2020\|EPI_ISL_778788\|20 | ···················· | ···················· | ···················· | ···················· | ···················· | ···················· | |
| AB | 8705 | hCoV-19/England/QEUH-CAC37A/2020\|EPI_ISL_760686\|20 | ···················· | ···················· | ···················· | ···················· | ······T············· | ···················· | |
| AB | 8706 | hCoV-19/England/QEUH-CC393F/2020\|EPI_ISL_760687\|20 | ···················· | ···················· | ···················· | ···················· | ···················· | ···················· | |
| AB | 8707 | hCoV-19/England/ALDP-CBE5D9/2020\|EPI_ISL_760688\|20 | ···················· | ···················· | ···················· | ···················· | ···················· | ···················· | |
| AB | 8708 | hCoV-19/England/QEUH-CC3CDC/2020\|EPI_ISL_760689\|20 | ···················· | ···················· | ···················· | ···················· | ···················· | ···················· | |
| AB | 8709 | hCoV-19/England/QEUH-CC2C28/2020\|EPI_ISL_760690\|20 | ···················· | ···················· | ···················· | ···················· | ···················· | ···A················ | |
| AB | 8710 | hCoV-19/England/QEUH-CC3D33/2020\|EPI_ISL_760695\|20 | ···················· | ···················· | ···················· | ···················· | ······T············· | ···················· | |
| AB | 8711 | hCoV-19/England/QEUH-CC386F/2020\|EPI_ISL_760696\|20 | ···················· | ···················· | ···················· | ···················· | ···················· | ···················· | |
| AB | 8712 | hCoV-19/England/CAMC-C7098C/2020\|EPI_ISL_769033\|20 | ···················· | ···················· | ···················· | ···················· | ···················· | ···················· | |
| AB | 8713 | hCoV-19/England/CAMC-C77132/2020\|EPI_ISL_769036\|20 | ···················· | ···················· | ···················· | ···················· | ···················· | ···················· | |
| AB | 8714 | hCoV-19/England/CAMC-C7716F/2020\|EPI_ISL_769038\|20 | ···················· | ···················· | ···················· | ···················· | ···················· | ···················· | |
| AB | 8715 | hCoV-19/Costa Rica/INC-0161/2020\|EPI_ISL_770002\|20 | ···················· | ···················· | ···················· | ···················· | ···················· | ···················· | |
| AB | 8716 | hCoV-19/Costa Rica/INC-0173/2020\|EPI_ISL_770003\|20 | ···················· | ···················· | ···················· | ···················· | ···················· | ···················· | |
| AB | 8717 | hCoV-19/Costa Rica/INC-0159/2020\|EPI_ISL_770004\|20 | ···················· | ···················· | ···················· | ···················· | ···················· | ···················· | |
| AB | 8718 | hCoV-19/Costa Rica/INC-0185/2020\|EPI_ISL_770005\|20 | ···················· | ···················· | ···················· | ···················· | ···················· | ···················· | |
| AB | 8719 | hCoV-19/Costa Rica/INC-0142/2020\|EPI_ISL_770006\|20 | ···················· | ···················· | ···················· | ···················· | ···················· | ···················· | |
| AB | 8720 | hCoV-19/Costa Rica/INC-0157/2020\|EPI_ISL_770007\|20 | ···················· | ···················· | ···················· | ···················· | ···················· | ···················· | |
| AB | 8721 | hCoV-19/Costa Rica/INC-0168/2020\|EPI_ISL_770008\|20 | ···················· | ···················· | ···················· | ···················· | ···················· | ···················· | |
| AB | 8722 | hCoV-19/Costa Rica/INC-0140/2020\|EPI_ISL_770009\|20 | ···················· | ···················· | ···················· | ···················· | ···················· | ···················· | |
| AB | 8723 | hCoV-19/Costa Rica/INC-0141/2020\|EPI_ISL_770010\|20 | ···················· | ···················· | ···················· | ···················· | ···················· | ···················· | |
| AB | 8724 | hCoV-19/Costa Rica/INC-0145/2020\|EPI_ISL_770011\|20 | ···················· | ···················· | ···················· | ···················· | ···················· | ···················· | |
| AB | 8725 | hCoV-19/Costa Rica/INC-0149/2020\|EPI_ISL_770012\|20 | ···················· | ···················· | ···················· | ···················· | ···················· | ···················· | |
| AB | 8726 | hCoV-19/Costa Rica/INC-0177/2020\|EPI_ISL_770020\|20 | ···················· | ···················· | ···················· | ···················· | ···················· | ···················· | |
| AB | 8727 | hCoV-19/Costa Rica/INC-0181/2020\|EPI_ISL_770021\|20 | ···················· | ···················· | ···················· | ···················· | ···················· | ···················· | |
| AB | 8728 | hCoV-19/Costa Rica/INC-0183/2020\|EPI_ISL_770022\|20 | ···················· | ···················· | ···················· | ···················· | ···················· | ···················· | |
| AB | 8729 | hCoV-19/Costa Rica/INC-0184/2020\|EPI_ISL_770023\|20 | ···················· | ···················· | ···················· | ···················· | ···················· | ···················· | |
| AB | 8730 | hCoV-19/USA/CO-CDPHE-2100023678/2020\|EPI_ISL_77133 | ···················· | ···················· | ···················· | ···················· | ···················· | ···················· | |
| AB | 8731 | hCoV-19/USA/CO-CDPHE-2100026492/2020\|EPI_ISL_77133 | ···················· | ···················· | ···················· | ···················· | ···················· | ···················· | |
| AB | 8732 | hCoV-19/USA/CO-CDPHE-2100028361/2020\|EPI_ISL_77133 | ···················· | ···················· | ···················· | ···················· | ···················· | ···················· | |
| AB | 8733 | hCoV-19/USA/CO-CDPHE-2100028367/2020\|EPI_ISL_77133 | ···················· | ···················· | ···················· | ···················· | ···················· | ···················· | |
| AB | 8735 | hCoV-19/USA/CO-CDPHE-2100028879/2020\|EPI_ISL_77133 | ···················· | ···················· | ···················· | ···················· | ···················· | ···················· | |
| AB | 8736 | hCoV-19/USA/CO-CDPHE-2100030859/2020\|EPI_ISL_77133 | ···················· | ···················· | ···················· | ···················· | ···················· | ···················· | |
| AB | 8737 | hCoV-19/USA/CO-CDPHE-2100036095/2020\|EPI_ISL_77134 | ···················· | ···················· | ···················· | ···················· | ···················· | ···················· | |
| AB | 8738 | hCoV-19/USA/CO-CDPHE-2100036219/2020\|EPI_ISL_77134 | ···················· | ···················· | ···················· | ···················· | ···················· | ···················· | |
| AB | 8739 | hCoV-19/USA/CO-CDPHE-2100036488/2020\|EPI_ISL_77134 | ···················· | ···················· | ···················· | ···················· | ···················· | ···················· | |
| AB | 8740 | hCoV-19/USA/CO-CDPHE-2100036496/2020\|EPI_ISL_77134 | ···················· | ···················· | ···················· | ···················· | ···················· | ···················· | |
| AB | 8741 | hCoV-19/USA/CO-CDPHE-2100038259/2020\|EPI_ISL_77135 | ···················· | ···················· | ···················· | ···················· | ···················· | ···················· | |
| AB | 8742 | hCoV-19/USA/CO-CDPHE-2100039270/2020\|EPI_ISL_77135 | ···················· | ···················· | ···················· | ···················· | ···················· | ···················· | |
| AB | 8743 | hCoV-19/USA/CO-CDPHE-2100039385/2020\|EPI_ISL_77135 | ···················· | ···················· | ···················· | ···················· | ···················· | ···················· | |
| AB | 8744 | hCoV-19/USA/CO-CDPHE-2100041649/2020\|EPI_ISL_77135 | ···················· | ···················· | ···················· | ···················· | ···················· | ···················· | |
| AB | 8745 | hCoV-19/USA/CO-CDPHE-2100042669/2020\|EPI_ISL_77135 | ···················· | ···················· | ···················· | ···················· | ···················· | ···················· | |
| Y | 1 | hCoV-19/Wales/PHWC-49725F/2020\|EPI_ISL_726800\|2020 | ···················· | ···················· | ···················· | ···················· | ···················N | ···················· | 1 |
| Y | 2 | hCoV-19/Wales/PHWC-49726E/2020\|EPI_ISL_726801\|2020 | ···················· | ···················· | ···················· | ···················· | ···················· | ···················· | |
| Y | 3 | hCoV-19/Wales/PHWC-49727D/2020\|EPI_ISL_726802\|2020 | ···················· | ···················· | ···················· | ···················· | ···················· | ···················· | |
| Y | 4 | hCoV-19/Wales/PHWC-49728C/2020\|EPI_ISL_726803\|2020 | ···················· | ···················· | ···················· | ···················· | ···················· | ···················· | |
| Y | 5 | hCoV-19/Wales/PHWC-49729B/2020\|EPI_ISL_726804\|2020 | ···················· | ···················· | ···················· | ···················· | ···················· | ···················· | |
| Y | 6 | hCoV-19/Switzerland/GR-ETHZ-400293/2020\|EPI_ISL_72 | ···················· | ···················· | ···················· | ···················· | ···················· | ···················· | |
| Y | 7 | hCoV-19/Switzerland/SO-ETHZ-400294/2020\|EPI_ISL_72 | ···················· | ···················· | ···················· | ···················· | ···················· | ···················· | |
| Y | 8 | hCoV-19/Switzerland/SO-ETHZ-400296/2020\|EPI_ISL_72 | ···················· | ···················· | ···················· | ···················· | ···················· | ···················· | |
| Y | 9 | hCoV-19/Switzerland/SO-ETHZ-400298/2020\|EPI_ISL_72 | ···················· | ···················· | ···················· | ···················· | ···················· | ···················· | |
| Y | 10 | hCoV-19/Switzerland/ZH-ETHZ-400376/2020\|EPI_ISL_72 | ···················· | ···················· | ···················· | ···················· | ···················· | ···················· | |
| Y | 11 | hCoV-19/Switzerland/ZH-ETHZ-400379/2020\|EPI_ISL_72 | ···················· | ···················· | ···················· | ···················· | ···················· | ···················· | |
| Y | 12 | hCoV-19/Switzerland/ZH-ETHZ-400380/2020\|EPI_ISL_72 | ···················· | ···················· | ···················· | ···················· | ···················· | ···················· | |
| Y | 13 | hCoV-19/Switzerland/BL-ETHZ-400387/2020\|EPI_ISL_72 | ···················· | ···················· | ···················· | ···················· | ···················· | ···················· | |
| Y | 14 | hCoV-19/Switzerland/BE-ETHZ-390148/2020\|EPI_ISL_72 | ···················· | ···················· | ···················· | ···················· | ···················· | ···················· | |
| Y | 15 | hCoV-19/Switzerland/ZH-ETHZ-390422/2020\|EPI_ISL_72 | ···················· | ···················· | ···················· | ···················· | ···················· | ···················· | |
| Y | 16 | hCoV-19/Switzerland/NE-ETHZ-390423/2020\|EPI_ISL_72 | ···················· | ···················· | ···················· | ···················· | ···················· | ···················· | |
| Y | 17 | hCoV-19/Switzerland/ZH-ETHZ-390426/2020\|EPI_ISL_72 | ···················· | ···················· | ···················· | ···················· | ···················· | ···················· | |
| Y | 18 | hCoV-19/Switzerland/NE-ETHZ-390427/2020\|EPI_ISL_72 | ···················· | ···················· | ···················· | ···················· | ···················· | ···················· | |
| Y | 19 | hCoV-19/Switzerland/SG-ETHZ-390429/2020\|EPI_ISL_72 | ···················· | ···················· | ···················· | ···················· | ···················· | ···················· | |
| Y | 20 | hCoV-19/Switzerland/ZH-ETHZ-390430/2020\|EPI_ISL_72 | ···················· | ···················· | ···················· | ···················· | ···················· | ···················· | |
| Y | 21 | hCoV-19/Switzerland/SG-ETHZ-390431/2020\|EPI_ISL_72 | ···················· | ···················· | ···················· | ···················· | ···················· | ···················· | |
| Y | 22 | hCoV-19/Switzerland/SG-ETHZ-390432/2020\|EPI_ISL_72 | ···················· | ···················· | ···················· | ···················· | ···················· | ···················· | |
| Y | 23 | hCoV-19/Switzerland/BE-ETHZ-390434/2020\|EPI_ISL_72 | ···················· | ···················· | ···················· | ···················· | ···················· | ···················· | |
| Y | 24 | hCoV-19/Switzerland/ZH-ETHZ-390435/2020\|EPI_ISL_72 | ···················· | ···················· | ···················· | ···················· | ···················· | ···················· | |

| | | | | | | | |
|---|---|---|---|---|---|---|---|
| Y | 25 hCoV-19/Switzerland/SG-ETHZ-390436/2020\|EPI_ISL_72 | ···················· | ···················· | ···················· | ···················· | ···················· | ···················· |
| Y | 26 hCoV-19/Nigeria/BO-CV337/2020\|EPI_ISL_730004\|2020- | ···················· | ···················· | ···················· | ···················· | ···················· | ···················· |
| Y | 27 hCoV-19/Nigeria/BO-CV339/2020\|EPI_ISL_730006\|2020- | ···················· | ···················· | ···················· | ···················· | ···················· | ···················· |
| Y | 28 hCoV-19/Nigeria/BO-CV341/2020\|EPI_ISL_730007\|2020- | ···················· | ···················· | ···················· | ···················· | ···················· | ···················· |
| Y | 29 hCoV-19/Nigeria/BO-CV344/2020\|EPI_ISL_730009\|2020- | ···················· | ···················· | ···················· | ···················· | ···················· | ···················· |
| Y | 30 hCoV-19/Nigeria/BO-CV345/2020\|EPI_ISL_730010\|2020- | ···················· | ···················· | ···················· | ···················· | ···················· | ···················· |
| Y | 31 hCoV-19/Nigeria/BO-CV347/2020\|EPI_ISL_730012\|2020- | ···················· | ···················· | ···················· | ···················· | ···················· | ···················· |
| Y | 32 hCoV-19/Nigeria/BO-CV348/2020\|EPI_ISL_730013\|2020- | ···················· | ···················· | ···················· | ···················· | ···················· | ···················· |
| Y | 33 hCoV-19/Nigeria/BO-CV350/2020\|EPI_ISL_730015\|2020- | ···················· | ···················· | ···················· | ···················· | ···················· | ···················· |
| Y | 34 hCoV-19/Nigeria/BE-CV352/2020\|EPI_ISL_730017\|2020- | ···················· | ···················· | ···················· | ···················· | ···················· | ···················· |
| Y | 35 hCoV-19/Nigeria/FC-CV354/2020\|EPI_ISL_730019\|2020- | ···················· | ···················· | ···················· | ···················· | ···················· | ···················· |
| Y | 36 hCoV-19/Nigeria/FC-CV355/2020\|EPI_ISL_730020\|2020- | ···················· | ···················· | ···················· | ···················· | ···················· | ···················· |
| Y | 37 hCoV-19/Nigeria/FC-CV356/2020\|EPI_ISL_730021\|2020- | ···················· | ···················· | ···················· | ···················· | ···················· | ···················· |
| Y | 38 hCoV-19/Nigeria/FC-CV360/2020\|EPI_ISL_730024\|2020- | ···················· | ···················· | ···················· | ···················· | ···················· | ···················· |
| Y | 39 hCoV-19/England/ALDP-C394BB/2020\|EPI_ISL_731302\|20 | ···················· | ···················· | ···················· | ···················· | ···················· | ···················· |
| Y | 40 hCoV-19/England/QEUH-C31714/2020\|EPI_ISL_731305\|20 | ···················· | ···················· | ···················· | ···················· | ···················· | ···················· |
| Y | 41 hCoV-19/England/QEUH-C316F9/2020\|EPI_ISL_731308\|20 | ···················· | ···················· | ···················· | ···················· | ···················· | ···················· |
| Y | 42 hCoV-19/England/QEUH-C30B0A/2020\|EPI_ISL_731309\|20 | ···················· | ···················· | ···················· | ···················· | ···················· | ···················· |
| Y | 43 hCoV-19/England/ALDP-C39512/2020\|EPI_ISL_731312\|20 | ···················· | ···················· | ···················· | ···················· | ······T············· | ···················· |
| Y | 44 hCoV-19/England/ALDP-C39AB9/2020\|EPI_ISL_731313\|20 | ···················· | ···················· | ···················· | ···················· | ···················· | ···················· |
| Y | 45 hCoV-19/England/QEUH-C31301/2020\|EPI_ISL_731314\|20 | ···················· | ···················· | ···················· | ···················· | ···················· | ···················· |
| Y | 46 hCoV-19/England/ALDP-C394F7/2020\|EPI_ISL_731318\|20 | ···················· | ···················· | ···················· | ···················· | ···················· | ···················· |
| Y | 47 hCoV-19/England/QEUH-C310FB/2020\|EPI_ISL_731319\|20 | ···················· | ···················· | ···················· | ···················· | ···················· | ···················· |
| Y | 48 hCoV-19/England/QEUH-C35FAF/2020\|EPI_ISL_731321\|20 | ···················· | ···················· | ···················· | ···················· | ···················· | ···················· |
| Y | 49 hCoV-19/England/ALDP-C3912D/2020\|EPI_ISL_731323\|20 | ···················· | ···················· | ···················· | ···················· | ···················· | ···················· |
| Y | 50 hCoV-19/England/ALDP-C3A7B1/2020\|EPI_ISL_731325\|20 | ···················· | ···················· | ···················· | ···················· | ···················· | ···················· |
| Y | 51 hCoV-19/England/QEUH-C35FDC/2020\|EPI_ISL_731326\|20 | ···················· | ···················· | ···················· | ···················· | ···················· | ···················· |
| Y | 52 hCoV-19/England/ALDP-C3A63C/2020\|EPI_ISL_731329\|20 | ···················· | ···················· | ···················· | ···················· | ···················· | ···················· |
| Y | 53 hCoV-19/England/QEUH-C318A8/2020\|EPI_ISL_731333\|20 | ········T··········· | ···················· | ···················· | ···················· | ···················· | ···················· |
| Y | 54 hCoV-19/England/QEUH-C31680/2020\|EPI_ISL_731334\|20 | ···················· | ···················· | ···················· | ···················· | ···················· | ···················· |
| Y | 55 hCoV-19/England/ALDP-C39530/2020\|EPI_ISL_731339\|20 | ···················· | ···················· | ···················· | ···················· | ···················· | ···················· |
| Y | 56 hCoV-19/England/QEUH-C30A3A/2020\|EPI_ISL_731341\|20 | ···················· | ···················· | ···················· | ···················· | ···················· | ···················· |
| Y | 57 hCoV-19/England/ALDP-C3AD37/2020\|EPI_ISL_731342\|20 | ···················· | ···················· | ···················· | ···················· | ···················· | ···················· |
| Y | 58 hCoV-19/England/ALDP-C3914B/2020\|EPI_ISL_731343\|20 | ···················· | ···················· | ···················· | ···················· | ···················· | ···················· |
| Y | 59 hCoV-19/England/ALDP-C39AD7/2020\|EPI_ISL_731344\|20 | ···················· | ···················· | ···················· | ···················· | ···················· | ···················· |
| Y | 60 hCoV-19/England/QEUH-C31091/2020\|EPI_ISL_731345\|20 | ···················· | ···················· | ···················· | ···················· | ···················· | ···················· |
| Y | 61 hCoV-19/England/QEUH-C31073/2020\|EPI_ISL_731346\|20 | ···················· | ···················· | ···················· | ···················· | ···················· | ···················· |
| Y | 62 hCoV-19/England/QEUH-C3132F/2020\|EPI_ISL_731349\|20 | ···················· | ···················· | ···················· | ···················· | ···················· | ···················· |
| Y | 63 hCoV-19/England/ALDP-C391C3/2020\|EPI_ISL_731351\|20 | ···················· | ···················· | ···················· | ···················· | ···················· | ···················· |
| Y | 64 hCoV-19/England/ALDP-C3AD0A/2020\|EPI_ISL_731352\|20 | ···················· | ···················· | ···················· | ···················· | ···················· | ···················· |
| Y | 65 hCoV-19/England/QEUH-C316DB/2020\|EPI_ISL_731353\|20 | ···················· | ···················· | ···················· | ···················· | ···················· | ···················· |
| Y | 66 hCoV-19/England/QEUH-C315A1/2020\|EPI_ISL_731354\|20 | ···················· | ···················· | ···················· | ···················· | ···················· | ···················· |
| Y | 67 hCoV-19/England/ALDP-C3A854/2020\|EPI_ISL_731358\|20 | ···················· | ···················· | ···················· | ···················· | ···················· | ···················· |
| Y | 68 hCoV-19/England/QEUH-C3119E/2020\|EPI_ISL_731382\|20 | ···················· | ···················· | ···················· | ···················· | ···················· | ···················· |
| Y | 69 hCoV-19/England/ALDP-C39A6E/2020\|EPI_ISL_731384\|20 | ···················· | ···················· | ···················· | ···················· | ···················· | ···················· |
| Y | 70 hCoV-19/England/ALDP-C3A20B/2020\|EPI_ISL_731385\|20 | ···················· | ···················· | ···················· | ···················· | ···················· | ···················· |
| Y | 71 hCoV-19/England/QEUH-C30B28/2020\|EPI_ISL_731386\|20 | ···················· | ···················· | ···················· | ···················· | ···················· | ···················· |
| Y | 72 hCoV-19/England/ALDP-C3ACFE/2020\|EPI_ISL_731388\|20 | ···················· | ···················· | ···················· | ···················· | ···················· | ···················· |
| Y | 73 hCoV-19/England/ALDP-C39257/2020\|EPI_ISL_731389\|20 | ···················· | ···················· | ···················· | ···················· | ···················· | ···················· |
| Y | 74 hCoV-19/England/QEUH-C315CF/2020\|EPI_ISL_731390\|20 | ···················· | ···················· | ···················· | ···················· | ········G··········· | ···················· |
| Y | 75 hCoV-19/England/ALDP-C3AD46/2020\|EPI_ISL_731391\|20 | ···················· | ···················· | ···················· | ···················· | ···················· | ···················· |
| Y | 76 hCoV-19/England/QEUH-C35FBE/2020\|EPI_ISL_731392\|20 | ···················· | ···················· | ···················· | ···················· | ···················· | ···················· |
| Y | 77 hCoV-19/England/QEUH-C30A2B/2020\|EPI_ISL_731393\|20 | ···················· | ···················· | ···················· | ···················· | ···················· | ···················· |
| Y | 78 hCoV-19/England/QEUH-C30FFF/2020\|EPI_ISL_731398\|20 | ···················· | ···················· | ···················· | ···················· | ···················· | ···················· |
| Y | 79 hCoV-19/England/ALDP-C392C0/2020\|EPI_ISL_731399\|20 | ···················· | ···················· | ···················· | ···················· | ···················· | ···················· |
| Y | 80 hCoV-19/Finland/1CE2/2020\|EPI_ISL_732600\|2020-12-0 | ···················· | ···················· | ···················· | ···················· | ···················· | ···················· |
| Y | 81 hCoV-19/Finland/1cF10/2020\|EPI_ISL_732601\|2020-10- | ···················· | ·A·················· | ···················· | ···················· | ···················· | ···················· |
| Y | 82 hCoV-19/Finland/1cF9/2020\|EPI_ISL_732602\|2020-10-3 | ···················· | ·A·················· | ···················· | ···················· | ···················· | ···A················ |
| Y | 83 hCoV-19/Finland/1cG10/2020\|EPI_ISL_732603\|2020-10- | ···················· | ·A·················· | ···················· | ···················· | ···················· | ···················· |
| Y | 84 hCoV-19/Finland/1CH5/2020\|EPI_ISL_732604\|2020-12-0 | ···················· | ···················· | ···················· | ···················· | ···················· | ···················· |
| Y | 85 hCoV-19/Finland/29C11/2020\|EPI_ISL_732605\|2020-12- | ···················· | ·A·················· | ···················· | ···················· | ···················· | ···················· |
| Y | 86 hCoV-19/Finland/29E10/2020\|EPI_ISL_732606\|2020-12- | ···················· | ·A·················· | ···················· | ···················· | ···················· | ···················· |
| Y | 87 hCoV-19/Finland/29E5/2020\|EPI_ISL_732607\|2020-12-0 | ···················· | ···················· | ···················· | ···················· | ···················· | ···················· |
| Y | 88 hCoV-19/Finland/29E6/2020\|EPI_ISL_732608\|2020-12-0 | ···················· | ···················· | ···················· | ···················· | ···················· | ···················· |
| Y | 89 hCoV-19/Finland/29F7/2020\|EPI_ISL_732609\|2020-12-0 | ···················· | ·A·················· | ···················· | ···················· | ···················· | ···················· |
| Y | 90 hCoV-19/Finland/29H7/2020\|EPI_ISL_732610\|2020-12-0 | ···················· | ···················· | ···················· | ···················· | ···················· | ···················· |
| Y | 91 hCoV-19/Finland/2CD6/2020\|EPI_ISL_732611\|2020-12-0 | ···················· | ···················· | ···················· | ···················· | ···················· | ···················· |
| Y | 92 hCoV-19/Finland/3CB4/2020\|EPI_ISL_732612\|2020-12-0 | ···················· | ·A·················· | ···················· | ···················· | ···················· | ···················· |
| Y | 94 hCoV-19/Finland/4DD5/2020\|EPI_ISL_732614\|2020-12-0 | ···················· | ···················· | ···················· | ···················· | ···················· | ···················· |
| Y | 95 hCoV-19/Finland/4DG9/2020\|EPI_ISL_732615\|2020-12-0 | ···················· | ···················· | ···················· | ···················· | ···················· | ···················· |
| Y | 96 hCoV-19/Finland/6CA1/2020\|EPI_ISL_732616\|2020-12-0 | ···················· | ···················· | ···················· | ···················· | ···················· | ···················· |
| Y | 97 hCoV-19/Finland/6CB1/2020\|EPI_ISL_732617\|2020-12-0 | ···················· | ···················· | ···················· | ···················· | ···················· | ···················· |
| Y | 98 hCoV-19/Finland/7cB5/2020\|EPI_ISL_732618\|2020-10-3 | ···················· | ···················· | ···················· | ···················· | ···················· | ···················· |
| Y | 99 hCoV-19/Finland/8CA1/2020\|EPI_ISL_732619\|2020-12-0 | ···················· | ···················· | ···················· | ···················· | ···················· | ···················· |
| Y | 100 hCoV-19/Finland/8CB1/2020\|EPI_ISL_732620\|2020-12-0 | ···················· | ···················· | ···················· | ···················· | ···················· | ···················· |
| Y | 101 hCoV-19/Finland/9CA12/2020\|EPI_ISL_732621\|2020-12- | ···················· | ·A·················· | ···················· | ···················· | ···················· | ···················· |
| Y | 102 hCoV-19/Finland/9CB12/2020\|EPI_ISL_732622\|2020-12- | ···················· | ·A·················· | ···················· | ···················· | ···················· | ···················· |
| Y | 103 hCoV-19/Finland/9CD10/2020\|EPI_ISL_732623\|2020-12- | ···················· | ···················· | ···················· | ···················· | ···················· | ···················· |
| Y | 104 hCoV-19/Finland/9CH11/2020\|EPI_ISL_732624\|2020-12- | ···················· | ·A·················· | ···················· | ···················· | ···················· | ···················· |
| Y | 105 hCoV-19/Finland/S10CB1/2020\|EPI_ISL_732625\|2020-10 | ···················· | ·A·················· | ···················· | ···················· | ···················· | ···A················ |

```
Y    106 hCoV-19/Finland/S10CF2/2020|EPI_ISL_732626|2020-10    ····················  ·A··················  ·····················  ····················  ····················  ···A················
Y    107 hCoV-19/Finland/S11BD1/2020|EPI_ISL_732627|2020-09    ····················  ····················  ·····················  ····················  ····················  ····················
Y    108 hCoV-19/Finland/S11CA6/2020|EPI_ISL_732628|2020-10    ····················  ····················  ·····················  ····················  ····················  ····················
Y    109 hCoV-19/Finland/S11F9/2020|EPI_ISL_732629|2020-09-    ····················  ·A··················  ·····················  ····················  ····················  ···A················
Y    110 hCoV-19/Finland/S11G9/2020|EPI_ISL_732630|2020-09-    ····················  ····················  ·····················  ····················  ····················  ····················
Y    111 hCoV-19/Finland/S12BE11/2020|EPI_ISL_732631|2020-0    ····················  ·A··················  ·····················  ····················  ····················  ···A················
Y    112 hCoV-19/Finland/S12CA6/2020|EPI_ISL_732632|2020-10    ····················  ····················  ·····················  ····················  ····················  ····················
Y    113 hCoV-19/Finland/S12CF9/2020|EPI_ISL_732633|2020-10    ····················  ····················  ·····················  ····················  ····················  ···A················
Y    114 hCoV-19/Finland/S13CC9/2020|EPI_ISL_732634|2020-10    ····················  ····················  ·····················  ····················  ····················  ····················
Y    115 hCoV-19/Finland/S17B1/2020|EPI_ISL_732635|2020-09-    ····················  ·A··················  ·····················  ····················  ····················  ···A················
Y    116 hCoV-19/Finland/S1CD10/2020|EPI_ISL_732636|2020-10    ····················  ·A··················  ·····················  ····················  ····················  ···A················
Y    117 hCoV-19/Finland/S2CA3/2020|EPI_ISL_732637|2020-10-    ····················  ····················  ·····················  ····················  ····················  ····················
Y    118 hCoV-19/Finland/S2CB7/2020|EPI_ISL_732638|2020-10-    ····················  ·A··················  ·····················  ····················  ····················  ····················
Y    119 hCoV-19/Finland/S2CG5/2020|EPI_ISL_732639|2020-10-    ····················  ····················  ·····················  ····················  ····················  ····················
Y    120 hCoV-19/Finland/S2CG8/2020|EPI_ISL_732640|2020-10-    ····················  ·A··················  ·····················  ····················  ····················  ····················
Y    121 hCoV-19/Finland/S3CA12/2020|EPI_ISL_732641|2020-10    ····················  ·A··················  ·····················  ····················  ····················  ····················
Y    122 hCoV-19/Finland/S3CA5/2020|EPI_ISL_732642|2020-10-    ····················  ·A··················  ·····················  ····················  ····················  ···A················
Y    123 hCoV-19/Finland/S3CB11/2020|EPI_ISL_732643|2020-10    ····················  ·A··················  ·····················  ····················  ····················  ····················
Y    124 hCoV-19/Finland/S3CC11/2020|EPI_ISL_732644|2020-10    ····················  ·A··················  ·····················  ····················  ····················  ····················
Y    125 hCoV-19/Finland/S3CC12/2020|EPI_ISL_732645|2020-10    ····················  ····················  ·····················  ····················  ····················  ····················
Y    126 hCoV-19/Finland/S3CD11/2020|EPI_ISL_732646|2020-10    ····················  ·A··················  ·····················  ····················  ····················  ····················
Y    127 hCoV-19/Finland/S3CE1/2020|EPI_ISL_732647|2020-10-    ····················  ·A··················  ·····················  ····················  ····················  ····················
Y    128 hCoV-19/Finland/S3CE11/2020|EPI_ISL_732648|2020-10    ····················  ····················  ·····················  ····················  ····················  ····················
Y    129 hCoV-19/Finland/S3CG11/2020|EPI_ISL_732649|2020-10    ····················  ·A··················  ·····················  ····················  ····················  ····················
Y    130 hCoV-19/Finland/S3CG2/2020|EPI_ISL_732650|2020-10-    ····················  ·A··················  ·····················  ····················  ····················  ····················
Y    131 hCoV-19/Finland/S5CD7/2020|EPI_ISL_732651|2020-10-    ····················  ·A··················  ·····················  ····················  ····················  ···A················
Y    132 hCoV-19/Finland/S9BC3/2020|EPI_ISL_732652|2020-09-    ····················  ····················  ·····················  ····················  ····················  ····················
Y    133 hCoV-19/Finland/S9CB11/2020|EPI_ISL_732653|2020-10    ····················  ·A··················  ·····················  ····················  ····················  ····················
Y    134 hCoV-19/Sweden/20-53609/2020|EPI_ISL_732654|2020-1    ····················  ····················  ·····················  ····················  ····················  ····················
Y    135 hCoV-19/Sweden/20-53518/2020|EPI_ISL_732655|2020-1    ···········T········  ····················  ·····················  ····················  ····················  ····················
Y    136 hCoV-19/Sweden/20-53632/2020|EPI_ISL_732656|2020-1    ····················  ····················  ·····················  ····················  ····················  ····················
Y    137 hCoV-19/Sweden/20-53652/2020|EPI_ISL_732657|2020-1    ····················  ····················  ·····················  ····················  ····················  ····················
Y    138 hCoV-19/Germany/BY-IMB11525/2020|EPI_ISL_732658|20    ····················  ····················  ·····················  ····················  ····················  ····················
Y    139 hCoV-19/Russia/MOS-CRIE-13472416/2020|EPI_ISL_7326    ····················  ····················  ·····················  ····················  ····················  ····················
Y    140 hCoV-19/Russia/MOS-CRIE-13382502/2020|EPI_ISL_7326    ····················  ····················  ·····················  ····················  ····················  ····················
Y    141 hCoV-19/Russia/MOS-CRIE-13384051/2020|EPI_ISL_7326    ····················  ····················  ·····················  ····················  ····················  ····················
Y    142 hCoV-19/Russia/MOS-CRIE-13364310/2020|EPI_ISL_7326    ····················  ····················  ·····················  ····················  ····················  ····················
Y    143 hCoV-19/Russia/MOS-CRIE-13385391/2020|EPI_ISL_7326    ····················  ····················  ·····················  ····················  ····················  ····················
Y    144 hCoV-19/Russia/MOS-CRIE-13378928/2020|EPI_ISL_7326    ····················  ····················  ·····················  ····················  ····················  ····················
Y    145 hCoV-19/Russia/MOS-CRIE-13381937/2020|EPI_ISL_7326    ····················  ····················  ·····················  ····················  ····················  ····················
Y    146 hCoV-19/Russia/MOS-CRIE-13363841/2020|EPI_ISL_7326    ····················  ····················  ·········T···········  ····················  ····················  ····················
Y    147 hCoV-19/Russia/MOS-CRIE-13378561/2020|EPI_ISL_7326    ····················  ····················  ·····················  ····················  ····················  ····················
Y    148 hCoV-19/Russia/MOS-CRIE-13383434/2020|EPI_ISL_7326    ····················  ······A·············  ·····················  ····················  ····················  ····················
Y    149 hCoV-19/Russia/MOS-CRIE-13383425/2020|EPI_ISL_7326    ····················  ····················  ·····················  ····················  ····················  ····················
Y    150 hCoV-19/Russia/MOS-CRIE-13385401/2020|EPI_ISL_7326    ····················  ····················  ·····················  ····················  ····················  ····················
Y    151 hCoV-19/Russia/MOS-CRIE-13431175/2020|EPI_ISL_7326    ····················  ····················  ·····················  ····················  ····················  ····················
Y    152 hCoV-19/Russia/MOS-CRIE-13385721/2020|EPI_ISL_7326    ····················  ····················  ·····················  ····················  ····················  ····················
Y    153 hCoV-19/Russia/MOS-CRIE-13365319/2020|EPI_ISL_7326    ····················  ····················  ·····················  ····················  ····················  ····················
Y    154 hCoV-19/Russia/MOS-CRIE-13384448/2020|EPI_ISL_7326    ····················  ····················  ·····················  ····················  ····················  ····················
Y    155 hCoV-19/Russia/MOS-CRIE-13444338/2020|EPI_ISL_7326    ····················  ····················  ·····················  ····················  ····················  ····················
Y    156 hCoV-19/Russia/MOS-CRIE-13298544/2020|EPI_ISL_7326    ····················  ····················  ·········T···········  ····················  ····················  ····················
Y    157 hCoV-19/France/ARA-100319/2020|EPI_ISL_732677|2020    ····················  ····················  ·····················  ····················  ····················  ····················
Y    158 hCoV-19/France/ARA-100644/2020|EPI_ISL_732678|2020    ····················  ····················  ·····················  ····················  ····················  ····················
Y    159 hCoV-19/France/ARA-101085/2020|EPI_ISL_732679|2020    ····················  ····················  ·····················  ····················  ····················  ····················
Y    160 hCoV-19/France/ARA-101986/2020|EPI_ISL_732680|2020    ····················  ····················  ·····················  ····················  ····················  ····················
Y    161 hCoV-19/France/ARA-102363/2020|EPI_ISL_732681|2020    ····················  ····················  ·····················  ····················  ····················  ····················
Y    162 hCoV-19/France/ARA-102618/2020|EPI_ISL_732682|2020    ····················  ····················  ·····················  ····················  ····················  ····················
Y    163 hCoV-19/France/ARA-103896/2020|EPI_ISL_732683|2020    ····················  ····················  ·····················  ····················  ····················  ····················
Y    164 hCoV-19/France/ARA-106011/2020|EPI_ISL_732684|2020    ····················  ····················  ·····················  ····················  ····················  ····················
Y    165 hCoV-19/France/ARA-106089/2020|EPI_ISL_732685|2020    ····················  ····················  ·····················  ····················  ····················  ····················
Y    166 hCoV-19/France/ARA-107483/2020|EPI_ISL_732686|2020    ····················  ····················  ·····················  ····················  ······T·············  ····················
Y    167 hCoV-19/France/ARA-108320/2020|EPI_ISL_732687|2020    ····················  ····················  ·····················  ····················  ····················  ····················
Y    168 hCoV-19/France/ARA-108984/2020|EPI_ISL_732688|2020    ····················  ····················  ·····················  ····················  ····················  ····················
Y    169 hCoV-19/France/ARA-110756/2020|EPI_ISL_732689|2020    ····················  ····················  ·····················  ····················  ····················  ····················
Y    170 hCoV-19/England/MILK-C30539/2020|EPI_ISL_733900|20    ····················  ····················  ·····················  ····················  ····················  ····················
Y    171 hCoV-19/England/ALDP-C23CBC/2020|EPI_ISL_733901|20    ····················  ····················  ·····················  ····················  ····················  ····················
Y    172 hCoV-19/England/MILK-C2F6C1/2020|EPI_ISL_733902|20    ····················  ····················  ·····················  ····················  ····················  ····················
Y    173 hCoV-19/England/MILK-C2FC92/2020|EPI_ISL_733903|20    ····················  ····················  ·····················  ····················  ····················  ····················
Y    174 hCoV-19/England/MILK-C2FAA7/2020|EPI_ISL_733904|20    ····················  ····················  ·····················  ····················  ····················  ····················
Y    175 hCoV-19/England/CAMC-C2667F/2020|EPI_ISL_733906|20    ····················  ····················  ·····················  ····················  ····················  ····················
Y    176 hCoV-19/England/CAMC-C2C907/2020|EPI_ISL_733909|20    ····················  ·A··················  ·····················  ····················  ····················  ····················
Y    177 hCoV-19/England/CAMC-C266AC/2020|EPI_ISL_733910|20    ····················  ····················  ·····················  ····················  ····················  ····················
Y    178 hCoV-19/England/ALDP-C23074/2020|EPI_ISL_733912|20    ····················  ····················  ·····················  ····················  ····················  ····················
Y    179 hCoV-19/England/ALDP-C244A4/2020|EPI_ISL_733913|20    ····················  ····················  ···········Y·········  ····················  ····················  ····················
Y    180 hCoV-19/England/MILK-C2FC65/2020|EPI_ISL_733914|20    ····················  ····················  ·····················  ····················  ····················  ····················
Y    181 hCoV-19/England/QEUH-C33F29/2020|EPI_ISL_733916|20    ····················  ····················  ·····················  ····················  ····················  ····················
Y    182 hCoV-19/England/QEUH-C34535/2020|EPI_ISL_733918|20    ····················  ····················  ·····················  ····················  ····················  ····················
Y    183 hCoV-19/England/QEUH-C348F0/2020|EPI_ISL_733919|20    ····················  ····················  ·····················  ····················  ····················  ····················
Y    184 hCoV-19/England/QEUH-C344CF/2020|EPI_ISL_733920|20    ····················  ····················  ·····················  ····················  ····················  ····················
Y    185 hCoV-19/England/QEUH-C345DB/2020|EPI_ISL_733922|20    ····················  ····················  ·····················  ····················  ······T·············  ····················
```

| | | | | | | | | |
|---|---|---|---|---|---|---|---|---|
| Y | 186 | hCoV-19/England/QEUH-C3376D/2020\|EPI_ISL_733923\|20 | ···················· | ···················· | ···················· | ···················· | ···················· | ···················· |
| Y | 187 | hCoV-19/England/QEUH-C3427A/2020\|EPI_ISL_733924\|20 | ···················· | ···················· | ···················· | ···················· | ···················· | ···················· |
| Y | 188 | hCoV-19/England/QEUH-C3408F/2020\|EPI_ISL_733925\|20 | ···················· | ···················· | ···················· | ···················· | ·····T·············· | ···················· |
| Y | 189 | hCoV-19/England/QEUH-C342F2/2020\|EPI_ISL_733926\|20 | ···················· | ···················· | ···················· | ···················· | ···················· | ···················· |
| Y | 190 | hCoV-19/England/QEUH-C346BA/2020\|EPI_ISL_733927\|20 | ···················· | ···················· | ···················· | ···················· | ···················· | ···················· |
| Y | 191 | hCoV-19/England/QEUH-C343B3/2020\|EPI_ISL_733928\|20 | ···················· | ···················· | ···················· | ···················· | ···················· | ···················· |
| Y | 192 | hCoV-19/England/QEUH-C33F56/2020\|EPI_ISL_733929\|20 | ···················· | ···················· | ···················· | ···················· | ···················· | ···················· |
| Y | 193 | hCoV-19/England/QEUH-C3383D/2020\|EPI_ISL_733930\|20 | ···················· | ···················· | ···················· | ···················· | ···················· | ···················· |
| Y | 194 | hCoV-19/England/QEUH-C332D5/2020\|EPI_ISL_733931\|20 | ···················· | ···················· | ···················· | ···················· | ···················· | ···················· |
| Y | 195 | hCoV-19/England/QEUH-C3374F/2020\|EPI_ISL_733932\|20 | ···················· | ···················· | ···················· | ···················· | ···················· | ···················· |
| Y | 196 | hCoV-19/England/QEUH-C33CC8/2020\|EPI_ISL_733934\|20 | ···················· | ···················· | ···················· | ···················· | ···················· | ···················· |
| Y | 197 | hCoV-19/England/QEUH-C34869/2020\|EPI_ISL_733935\|20 | ···················· | ···················· | ···················· | ···················· | ···················· | ···················· |
| Y | 198 | hCoV-19/England/QEUH-C3473F/2020\|EPI_ISL_733937\|20 | ···················· | ···················· | ···················· | ···················· | ···················· | ···················· |
| Y | 199 | hCoV-19/England/QEUH-C3392B/2020\|EPI_ISL_733939\|20 | ···················· | ···················· | ···················· | ···················· | ···················· | ···················· |
| Y | 200 | hCoV-19/England/QEUH-C33ED1/2020\|EPI_ISL_733940\|20 | ···················· | ···················· | ···················· | ···················· | ···················· | ···················· |
| Y | 201 | hCoV-19/England/QEUH-C343A4/2020\|EPI_ISL_733941\|20 | ···················· | ···················· | ···················· | ···················· | ···················· | ···················· |
| Y | 202 | hCoV-19/England/QEUH-C33C5F/2020\|EPI_ISL_733943\|20 | ···················· | ···················· | ···················· | ···················· | ···················· | ···················· |
| Y | 203 | hCoV-19/England/QEUH-C34799/2020\|EPI_ISL_733944\|20 | ···················· | ···················· | ···················· | ···················· | ···················· | ···················· |
| Y | 204 | hCoV-19/England/QEUH-C33C6E/2020\|EPI_ISL_733946\|20 | ···················· | ···················· | ···················· | ···················· | ···················· | ···················· |
| Y | 205 | hCoV-19/England/QEUH-C347D5/2020\|EPI_ISL_733948\|20 | ···················· | ···················· | ···················· | ···················· | ···················· | ···················· |
| Y | 206 | hCoV-19/England/QEUH-C3441A/2020\|EPI_ISL_733950\|20 | ···················· | ···················· | ···················· | ···················· | ···················· | ···················· |
| Y | 207 | hCoV-19/England/QEUH-C3317E/2020\|EPI_ISL_733951\|20 | ···················· | ···················· | ···················· | ···················· | ···················· | ···················· |
| Y | 208 | hCoV-19/England/QEUH-C33FFC/2020\|EPI_ISL_733953\|20 | ···················· | ···················· | ···················· | ···················· | ···················· | ···················· |
| Y | 209 | hCoV-19/England/QEUH-C3391C/2020\|EPI_ISL_733954\|20 | ···················· | ···················· | ···················· | ···················· | ···················· | ···················· |
| Y | 210 | hCoV-19/England/QEUH-C340BC/2020\|EPI_ISL_733955\|20 | ···················· | ···················· | ···················· | ···················· | ···················· | ···················· |
| Y | 211 | hCoV-19/England/QEUH-C32E0F/2020\|EPI_ISL_733956\|20 | ···················· | ···················· | ···················· | ···················· | ···················· | ···················· |
| Y | 212 | hCoV-19/England/QEUH-C33B34/2020\|EPI_ISL_733957\|20 | ···················· | ···················· | ···················· | ···················· | ···················· | ···················· |
| Y | 213 | hCoV-19/England/QEUH-C34650/2020\|EPI_ISL_733959\|20 | ···················· | ···················· | ···················· | ···················· | ···················· | ···················· |
| Y | 214 | hCoV-19/England/QEUH-C33CD7/2020\|EPI_ISL_733961\|20 | ···················· | ···················· | ···················· | ···················· | ···················· | ···················· |
| Y | 215 | hCoV-19/England/QEUH-C33B52/2020\|EPI_ISL_733962\|20 | ···················· | ···················· | ···················· | ···················· | ···················· | ···················· |
| Y | 216 | hCoV-19/England/QEUH-C331BA/2020\|EPI_ISL_733964\|20 | ···················· | ···················· | ···················· | ···················· | ···················· | ···················· |
| Y | 217 | hCoV-19/England/QEUH-C345CC/2020\|EPI_ISL_733967\|20 | ···················· | ···················· | ···················· | ···················· | ···················· | ···················· |
| Y | 218 | hCoV-19/England/QEUH-C3334B/2020\|EPI_ISL_733968\|20 | ···················· | ···················· | ···················· | ···················· | ···················· | ···················· |
| Y | 219 | hCoV-19/England/QEUH-C34474/2020\|EPI_ISL_733969\|20 | ···················· | ···················· | ···················· | ···················· | ···················· | ···················· |
| Y | 220 | hCoV-19/England/QEUH-C33B07/2020\|EPI_ISL_733970\|20 | ···················· | ···················· | ···················· | ···················· | ···················· | ···················· |
| Y | 221 | hCoV-19/England/QEUH-C338C4/2020\|EPI_ISL_733971\|20 | ···················· | ···················· | ···················· | ···················· | ···················· | ···················· |
| Y | 222 | hCoV-19/England/QEUH-C341AA/2020\|EPI_ISL_733972\|20 | ···················· | ···················· | ···················· | ···················· | ·····T·············· | ···················· |
| Y | 223 | hCoV-19/England/QEUH-C32EF0/2020\|EPI_ISL_733973\|20 | ···················· | ···················· | ···················· | ···················· | ···················· | ···················· |
| Y | 224 | hCoV-19/England/QEUH-C342C5/2020\|EPI_ISL_733974\|20 | ···················· | ···················· | ···················· | ···················· | ···················· | ···················· |
| Y | 225 | hCoV-19/England/QEUH-C345EA/2020\|EPI_ISL_733975\|20 | ···················· | ···················· | ···················· | ···················· | ···················· | ···················· |
| Y | 226 | hCoV-19/England/QEUH-C3482D/2020\|EPI_ISL_733976\|20 | ···················· | ···················· | ···················· | ···················· | ···················· | ···················· |
| Y | 227 | hCoV-19/England/QEUH-C3426B/2020\|EPI_ISL_733978\|20 | ···················· | ···················· | ···················· | ···················· | ···················· | ···················· |
| Y | 228 | hCoV-19/England/QEUH-C33FED/2020\|EPI_ISL_733979\|20 | ···················· | ···················· | ···················· | ···················· | ···················· | ···················· |
| Y | 229 | hCoV-19/England/QEUH-C3485A/2020\|EPI_ISL_733980\|20 | ···················· | ···················· | ···················· | ···················· | ···················· | ···················· |
| Y | 230 | hCoV-19/England/QEUH-C348A5/2020\|EPI_ISL_733981\|20 | ···················· | ···················· | ···················· | ···················· | ···················· | ···················· |
| Y | 231 | hCoV-19/England/QEUH-C34526/2020\|EPI_ISL_733983\|20 | ···················· | ···················· | ···················· | ···················· | ···················· | ···················· |
| Y | 232 | hCoV-19/England/QEUH-C331F6/2020\|EPI_ISL_733984\|20 | ···················· | ···················· | ···················· | ···················· | ···················· | ···················· |
| Y | 233 | hCoV-19/England/QEUH-C331AB/2020\|EPI_ISL_733985\|20 | ···················· | ···················· | ···················· | ···················· | ·····T·············· | ···················· |
| Y | 234 | hCoV-19/England/QEUH-C32F1B/2020\|EPI_ISL_733986\|20 | ···················· | ···················· | ···················· | ···················· | ···················· | ···················· |
| Y | 235 | hCoV-19/England/QEUH-C33202/2020\|EPI_ISL_733987\|20 | ···················· | ···················· | ···················· | ···················· | ···················· | ···················· |
| Y | 236 | hCoV-19/England/QEUH-C32E5A/2020\|EPI_ISL_733988\|20 | ···················· | ···················· | ···················· | ···················· | ···················· | ···················· |
| Y | 237 | hCoV-19/England/QEUH-C3475D/2020\|EPI_ISL_733989\|20 | ···················· | ···················· | ···················· | ···················· | ···················· | ···················· |
| Y | 238 | hCoV-19/England/QEUH-C32E69/2020\|EPI_ISL_733990\|20 | ···················· | ···················· | ···················· | ···················· | ···················· | ···················· |
| Y | 239 | hCoV-19/England/QEUH-C32F39/2020\|EPI_ISL_733991\|20 | ···················· | ···················· | ···················· | ···················· | ···················· | ···················· |
| Y | 240 | hCoV-19/England/QEUH-C348B4/2020\|EPI_ISL_733996\|20 | ···················· | ···················· | ···················· | ···················· | ···················· | ···················· |
| Y | 241 | hCoV-19/England/QEUH-C34544/2020\|EPI_ISL_733997\|20 | ···················· | ···················· | ···················· | ···················· | ···················· | ···················· |
| Y | 242 | hCoV-19/England/QEUH-C340F8/2020\|EPI_ISL_733998\|20 | ···················· | ···················· | ···················· | ···················· | ···················· | ···················· |
| Y | 243 | hCoV-19/England/QEUH-C3490C/2020\|EPI_ISL_733999\|20 | ···················· | ···················· | ···················· | ···················· | ···················· | ···················· |
| Y | 244 | hCoV-19/England/ALDP-C5A047/2020\|EPI_ISL_736173\|20 | ···················· | ···················· | ···················· | ···················· | ···················· | ···················· |
| Y | 245 | hCoV-19/England/QEUH-C5D4A2/2020\|EPI_ISL_736182\|20 | ···················· | ···················· | ···················· | ···················· | ···················· | ···················· |
| Y | 246 | hCoV-19/England/QEUH-C5BD03/2020\|EPI_ISL_736183\|20 | ···················· | ···················· | ···················· | ···················· | ···················· | ···················· |
| Y | 247 | hCoV-19/England/QEUH-C5D527/2020\|EPI_ISL_736184\|20 | ···················· | ···················· | ···················· | ···················· | ···················· | ···················· |
| Y | 248 | hCoV-19/England/ALDP-C58C18/2020\|EPI_ISL_736187\|20 | ···················· | ···················· | ···················· | ···················· | ···················· | ···················· |
| Y | 249 | hCoV-19/England/ALDP-C57F4D/2020\|EPI_ISL_736189\|20 | ···················· | ···················· | ···················· | ···················· | ···················· | ···················· |
| Y | 250 | hCoV-19/England/ALDP-C58191/2020\|EPI_ISL_736192\|20 | ···················· | ···················· | ···················· | ···················· | ···················· | ···················· |
| Y | 251 | hCoV-19/England/ALDP-C57CCE/2020\|EPI_ISL_736193\|20 | ···················· | ···················· | ···················· | ···················· | ···················· | ···················· |
| Y | 252 | hCoV-19/England/ALDP-C581A0/2020\|EPI_ISL_736194\|20 | ···················· | ···················· | ···················· | ···················· | ···················· | ···················· |
| Y | 253 | hCoV-19/England/ALDP-C5810A/2020\|EPI_ISL_736196\|20 | ···················· | ···················· | ···················· | ···················· | ···················· | ···················· |
| Y | 254 | hCoV-19/England/ALDP-C58CAF/2020\|EPI_ISL_736197\|20 | ···················· | ···················· | ···················· | ···················· | ···················· | ···················· |
| Y | 255 | hCoV-19/England/ALDP-C59FF0/2020\|EPI_ISL_736198\|20 | ···················· | ···················· | ···················· | ···················· | ···················· | ···················· |
| Y | 256 | hCoV-19/England/ALDP-C58577/2020\|EPI_ISL_736199\|20 | ···················· | ···················· | ···················· | ···················· | ···················· | ···················· |
| Y | 257 | hCoV-19/England/NORW-FA3D3/2020\|EPI_ISL_725501\|202 | ···················· | ···················· | ···················· | ···················· | ···················· | ···················· |
| Y | 258 | hCoV-19/England/NORW-FA3E2/2020\|EPI_ISL_725502\|202 | ···················· | ···················· | ···················· | ···················· | ···················· | ···················· |
| Y | 259 | hCoV-19/England/NORW-FA449/2020\|EPI_ISL_725505\|202 | ···················· | ···················· | ···················· | ···················· | ···················· | ···················· |
| Y | 260 | hCoV-19/England/NORW-FA485/2020\|EPI_ISL_725507\|202 | ···················· | ···················· | ···················· | ···················· | ···················· | ···················· |
| Y | 261 | hCoV-19/England/NORW-FA528/2020\|EPI_ISL_725510\|202 | ···················· | ···················· | ···················· | ···················· | ···················· | ···················· |
| Y | 262 | hCoV-19/England/NORW-FA537/2020\|EPI_ISL_725511\|202 | ···················· | ···················· | ···················· | ···················· | ···················· | ···················· |
| Y | 263 | hCoV-19/England/NORW-FA5DD/2020\|EPI_ISL_725516\|202 | ···················· | ···················· | ···················· | ···················· | ···················· | ···················· |
| Y | 264 | hCoV-19/England/NOTT-11E305/2020\|EPI_ISL_725525\|20 | ···················· | ···················· | ···················· | ···················· | ···················· | ···················· |
| Y | 265 | hCoV-19/England/NOTT-11E745/2020\|EPI_ISL_725534\|20 | ···················· | ···················· | ···················· | ···················· | ···················· | ···················· |

```
Y  266 hCoV-19/England/NOTT-11E754/2020|EPI_ISL_725535|20   ····················  ····················  ·····················  ····················  ····················  ····················
Y  267 hCoV-19/England/NOTT-11E763/2020|EPI_ISL_725536|20   ····················  ····················  ·····················  ····················  ····················  ····················
Y  268 hCoV-19/England/NOTT-11E772/2020|EPI_ISL_725537|20   ····················  ····················  ·····················  ····················  ····················  ····················
Y  269 hCoV-19/England/NOTT-11EF7A/2020|EPI_ISL_725538|20   ····················  ····················  ·····················  ····················  ····················  ····················
Y  270 hCoV-19/England/NOTT-11F559/2020|EPI_ISL_725541|20   ····················  ·A··················  ·····················  ····················  ····················  ····················
Y  271 hCoV-19/England/NOTT-11F5B3/2020|EPI_ISL_725544|20   ····················  ····················  ·····················  ····················  ····················  ····················
Y  272 hCoV-19/England/NOTT-11F5D1/2020|EPI_ISL_725545|20   ····················  ····················  ·····················  ····················  ····················  ····················
Y  273 hCoV-19/England/NOTT-11F5FF/2020|EPI_ISL_725546|20   ····················  ·A··················  ·····················  ····················  ····················  ····················
Y  274 hCoV-19/England/NOTT-11F629/2020|EPI_ISL_725548|20   ····················  ····················  ·····················  ····················  ····················  ····················
Y  275 hCoV-19/England/NOTT-11F638/2020|EPI_ISL_725549|20   ····················  ····················  ·····················  ····················  ····················  ····················
Y  276 hCoV-19/England/NOTT-11F656/2020|EPI_ISL_725550|20   ····················  ····················  ·····················  ····················  ····················  ····················
Y  277 hCoV-19/England/NOTT-11F692/2020|EPI_ISL_725552|20   ····················  ····················  ·····················  ····················  ····················  ····················
Y  278 hCoV-19/England/NOTT-11F6A1/2020|EPI_ISL_725553|20   ····················  ····················  ·····················  ····················  ····················  ····················
Y  279 hCoV-19/England/NOTT-11F6CF/2020|EPI_ISL_725555|20   ····················  ····················  ·····················  ····················  ····················  ····················
Y  280 hCoV-19/England/NOTT-11F6DE/2020|EPI_ISL_725556|20   ····················  ····················  ·····················  ····················  ····················  ····················
Y  281 hCoV-19/England/NOTT-11F6FC/2020|EPI_ISL_725558|20   ····················  ····················  ·····················  ····················  ····················  ····················
Y  282 hCoV-19/England/NOTT-11F717/2020|EPI_ISL_725559|20   ····················  ····················  ·····················  ····················  ····················  ····················
Y  283 hCoV-19/England/NOTT-11F726/2020|EPI_ISL_725560|20   ····················  ····················  ·····················  ····················  ····················  ····················
Y  284 hCoV-19/England/NOTT-11F762/2020|EPI_ISL_725563|20   ····················  ····················  ·····················  ····················  ····················  ····················
Y  285 hCoV-19/England/NOTT-11F771/2020|EPI_ISL_725564|20   ····················  ····················  ·····················  ····················  ····················  ····················
Y  286 hCoV-19/England/NOTT-11F7BD/2020|EPI_ISL_725568|20   ····················  ····················  ·····················  ····················  ····················  ····················
Y  287 hCoV-19/England/NOTT-11F7CC/2020|EPI_ISL_725569|20   ····················  ····················  ·····················  ····················  ····················  ····················
Y  288 hCoV-19/England/NOTT-11F7DB/2020|EPI_ISL_725570|20   ····················  ····················  ·····················  ····················  ····················  ····················
Y  289 hCoV-19/England/NOTT-11F7EA/2020|EPI_ISL_725571|20   ····················  ····················  ·····················  ····················  ····················  ····················
Y  290 hCoV-19/England/NOTT-11F805/2020|EPI_ISL_725573|20   ····················  ····················  ·····················  ····················  ····················  ····················
Y  291 hCoV-19/England/NOTT-11F814/2020|EPI_ISL_725574|20   ····················  ····················  ·····················  ····················  ····················  ····················
Y  292 hCoV-19/England/NOTT-11F86F/2020|EPI_ISL_725577|20   ····················  ····················  ·····················  ····················  ····················  ····················
Y  293 hCoV-19/England/NOTT-11F8C9/2020|EPI_ISL_725581|20   ····················  ····················  ·····················  ····················  ····················  ····················
Y  294 hCoV-19/England/NOTT-11F8F6/2020|EPI_ISL_725583|20   ····················  ····················  ·····················  ····················  ····················  ····················
Y  295 hCoV-19/England/NOTT-11F920/2020|EPI_ISL_725586|20   ····················  ····················  ·····················  ····················  ····················  ····················
Y  296 hCoV-19/England/NOTT-11F93F/2020|EPI_ISL_725587|20   ····················  ····················  ·····················  ····················  ····················  ····················
Y  297 hCoV-19/England/NOTT-11F94E/2020|EPI_ISL_725588|20   ····················  ····················  ·····················  ····················  ····················  ····················
Y  298 hCoV-19/England/NOTT-11F95D/2020|EPI_ISL_725589|20   ····················  ····················  ·····················  ····················  ····················  ····················
Y  299 hCoV-19/England/NOTT-11F98A/2020|EPI_ISL_725592|20   ····················  ····················  ·····················  ····················  ····················  ····················
Y  300 hCoV-19/England/NOTT-11F9B7/2020|EPI_ISL_725594|20   ····················  ····················  ·····················  ····················  ····················  ····················
Y  301 hCoV-19/Wales/PHWC-4972AA/2020|EPI_ISL_726805|2020   ····················  ····················  ·····················  ····················  ····················  NN··················
Y  302 hCoV-19/Wales/PHWC-4972B9/2020|EPI_ISL_726806|2020   ····················  ····················  ·····················  ····················  ····················  ····················
Y  303 hCoV-19/Wales/PHWC-4972C8/2020|EPI_ISL_726807|2020   ····················  ····················  ·····················  ····················  ····················  ····················
Y  304 hCoV-19/Wales/PHWC-4972D7/2020|EPI_ISL_726808|2020   ····················  ····················  ·····················  ····················  ····················  ····················
Y  305 hCoV-19/Wales/PHWC-4972E6/2020|EPI_ISL_726809|2020   ····················  ····················  ·····················  ····················  ····················  ····················
Y  306 hCoV-19/Wales/PHWC-4972F5/2020|EPI_ISL_726810|2020   ····················  ····················  ·····················  ····················  ····················  ····················
Y  307 hCoV-19/Wales/PHWC-497301/2020|EPI_ISL_726811|2020   ····················  ····················  ·····················  ····················  ····················  ····················
Y  311 hCoV-19/Wales/PHWC-49734D/2020|EPI_ISL_726815|2020   ····················  ····················  ·····················  ····················  ····················  ····················
Y  312 hCoV-19/Wales/PHWC-49735C/2020|EPI_ISL_726816|2020   ····················  ····················  ·····················  ····················  ····················  ····················
Y  313 hCoV-19/Wales/PHWC-49736B/2020|EPI_ISL_726817|2020   ····················  ····················  ·····················  ····················  ····················  ····················
Y  314 hCoV-19/Wales/PHWC-49737A/2020|EPI_ISL_726818|2020   ····················  ····················  ·····················  ····················  ····················  ····················
Y  315 hCoV-19/Wales/PHWC-497389/2020|EPI_ISL_726819|2020   ····················  ····················  ·····················  ····················  ····················  ····················
Y  316 hCoV-19/Wales/PHWC-497398/2020|EPI_ISL_726820|2020   ····················  ····················  ·····················  ····················  ····················  ····················
Y  317 hCoV-19/Wales/PHWC-4973A7/2020|EPI_ISL_726821|2020   ····················  ····················  ·····················  ····················  ····················  ····················
Y  318 hCoV-19/Wales/PHWC-4973B6/2020|EPI_ISL_726822|2020   ····················  ····················  ·····················  ····················  ····················  ····················
Y  319 hCoV-19/Wales/PHWC-4973C5/2020|EPI_ISL_726823|2020   ····················  ····················  ·····················  ····················  ····················  ····················
Y  320 hCoV-19/Wales/PHWC-4973D4/2020|EPI_ISL_726824|2020   ····················  ····················  ·····················  ····················  ····················  ····················
Y  321 hCoV-19/Wales/PHWC-4973E3/2020|EPI_ISL_726825|2020   ····················  ····················  ·····················  ····················  ····················  ····················
Y  322 hCoV-19/Wales/PHWC-4973F2/2020|EPI_ISL_726826|2020   ····················  ····················  ·····················  ····················  ····················  ····················
Y  324 hCoV-19/Wales/PHWC-49741D/2020|EPI_ISL_726828|2020   ····················  ····················  ·····················  ····················  ····················  ····················
Y  326 hCoV-19/Wales/PHWC-49743B/2020|EPI_ISL_726830|2020   ····················  ····················  ·····················  ····················  ····················  ····················
Y  327 hCoV-19/Wales/PHWC-49744A/2020|EPI_ISL_726831|2020   ····················  ····················  ·····················  ····················  ····················  ····················
Y  328 hCoV-19/Wales/PHWC-497459/2020|EPI_ISL_726832|2020   ····················  ····················  ·····················  ····················  ····················  ····················
Y  329 hCoV-19/Wales/PHWC-497486/2020|EPI_ISL_726833|2020   ····················  ····················  ·····················  ····················  ····················  ····················
Y  330 hCoV-19/Wales/PHWC-497495/2020|EPI_ISL_726834|2020   ····················  ····················  ·····················  ····················  NNN·················  ····················   1
Y  331 hCoV-19/Wales/PHWC-4974A4/2020|EPI_ISL_726835|2020   ····················  ····················  ·····················  ····················  ····················  ····················
Y  332 hCoV-19/Wales/PHWC-4974B3/2020|EPI_ISL_726836|2020   ····················  ····················  ·····················  ····················  ····················  ····················
Y  333 hCoV-19/Wales/PHWC-4974C2/2020|EPI_ISL_726837|2020   ····················  ······A·············  ·····················  ····················  ····················  ····················
Y  334 hCoV-19/Wales/PHWC-4974D1/2020|EPI_ISL_726838|2020   ····················  ····················  ·····················  ····················  ····················  ····················
Y  335 hCoV-19/Wales/PHWC-4974E0/2020|EPI_ISL_726839|2020   ····················  ····················  ·····················  ····················  ····················  ····················
Y  336 hCoV-19/Wales/PHWC-4974FF/2020|EPI_ISL_726840|2020   ····················  ····················  ·····················  ····················  ····················  ····················
Y  337 hCoV-19/Wales/PHWC-49750B/2020|EPI_ISL_726841|2020   ····················  ····················  ·····················  ····················  ····················  ····················
Y  338 hCoV-19/Wales/PHWC-49751A/2020|EPI_ISL_726842|2020   ····················  ····················  ·····················  ····················  ····················  ····················
Y  339 hCoV-19/Wales/PHWC-497529/2020|EPI_ISL_726843|2020   ····················  ····················  ·····················  ····················  ····················  ····················
Y  340 hCoV-19/Wales/PHWC-497547/2020|EPI_ISL_726845|2020   ····················  ····················  ·····················  ····················  ····················  ····················
Y  341 hCoV-19/Wales/PHWC-497556/2020|EPI_ISL_726846|2020   ····················  ····················  ·····················  ····················  ····················  ····················
Y  343 hCoV-19/Wales/PHWC-497574/2020|EPI_ISL_726848|2020   ····················  ····················  ·····················  ····················  ····················  ····················
Y  345 hCoV-19/Wales/PHWC-497592/2020|EPI_ISL_726850|2020   ····················  ····················  ·········A···········  ····················  ····················  ····················
Y  346 hCoV-19/Wales/PHWC-4975A1/2020|EPI_ISL_726851|2020   ····················  ····················  ·····················  ····················  ···T················  ····················
Y  347 hCoV-19/Wales/PHWC-4975CF/2020|EPI_ISL_726852|2020   ····················  ····················  ·····················  ····················  ····················  ····················
Y  349 hCoV-19/Wales/PHWC-4975FC/2020|EPI_ISL_726855|2020   ····················  ····················  ·····················  ····················  ····················  ····················
Y  350 hCoV-19/Wales/PHWC-497608/2020|EPI_ISL_726856|2020   ····················  ····················  ·····················  ····················  ····················  ····················
Y  351 hCoV-19/Wales/PHWC-497635/2020|EPI_ISL_726859|2020   ····················  ····················  ·····················  ····················  ····················  ····················
Y  353 hCoV-19/Wales/PHWC-497680/2020|EPI_ISL_726864|2020   ····················  ····················  ·····················  ····················  ····················  ····················
Y  354 hCoV-19/Wales/PHWC-49769F/2020|EPI_ISL_726865|2020   ····················  ····················  ·····················  ····················  ····················  ····················
```

```
Y 355 hCoV-19/Wales/PHWC-4976BD/2020|EPI_ISL_726866|2020 ···················· ···················· ···················· ···················· ···················· ····················
Y 356 hCoV-19/Wales/PHWC-4976CC/2020|EPI_ISL_726867|2020 ···················· ···················· ···················· ···················· ···················· ····················
Y 357 hCoV-19/Wales/PHWC-4976EA/2020|EPI_ISL_726869|2020 ···················· ···················· ···················· ···················· ···················· ····················
Y 358 hCoV-19/Wales/PHWC-4976F9/2020|EPI_ISL_726870|2020 ···················· ···················· ···················· ···················· ···················· ····················
Y 359 hCoV-19/Wales/PHWC-497705/2020|EPI_ISL_726871|2020 ···················· ···················· ···················· ···················· ···················· ····················
Y 360 hCoV-19/Wales/PHWC-497723/2020|EPI_ISL_726872|2020 ···················· ···················· ···················· ···················· ···················· ····················
Y 361 hCoV-19/Wales/PHWC-497732/2020|EPI_ISL_726873|2020 ···················· ···················· ···················· ···················· ···················· ····················
Y 362 hCoV-19/Wales/PHWC-497750/2020|EPI_ISL_726875|2020 ···················· ···················· ···················· ···················· ···················· ····················
Y 364 hCoV-19/Wales/PHWC-49777E/2020|EPI_ISL_726877|2020 ···················· ···················· ···················· ···················· ···················· ····················
Y 365 hCoV-19/Wales/PHWC-49778D/2020|EPI_ISL_726878|2020 ···················· ···················· ···················· ···················· ···················· ····················
Y 366 hCoV-19/Wales/PHWC-49779C/2020|EPI_ISL_726879|2020 ···················· ···················· ···················· ···················· ···················· ····················
Y 367 hCoV-19/Wales/PHWC-4977AB/2020|EPI_ISL_726880|2020 ···················· ···················· ···················· ···················· ···················· ····················
Y 369 hCoV-19/Wales/PHWC-4977C9/2020|EPI_ISL_726882|2020 ···················· ···················· ···················· ···················· ···················· ····················
Y 370 hCoV-19/Wales/PHWC-4977D8/2020|EPI_ISL_726883|2020 ···················· ···················· ···················· ···················· ···················· ····················
Y 371 hCoV-19/Wales/PHWC-4977E7/2020|EPI_ISL_726884|2020 ···················· ···················· ···················· ···················· ···················· ····················
Y 372 hCoV-19/Wales/PHWC-4977F6/2020|EPI_ISL_726885|2020 ···················· ···················· ···················· ···················· ···················· ····················
Y 373 hCoV-19/Wales/PHWC-497802/2020|EPI_ISL_726886|2020 ···················· ···················· ···················· ···················· ···················· ····················
Y 374 hCoV-19/Wales/PHWC-497811/2020|EPI_ISL_726887|2020 ···················· ···················· ···················· ···················· ···················· ····················
Y 375 hCoV-19/Wales/PHWC-497820/2020|EPI_ISL_726888|2020 ···················· ···················· ···················· ···················· ···················· ····················
Y 376 hCoV-19/Wales/PHWC-49783F/2020|EPI_ISL_726889|2020 ···················· ···················· ···················· ···················· ···················· ····················
Y 377 hCoV-19/Wales/PHWC-49785D/2020|EPI_ISL_726891|2020 ···················· ···················· ···················· ···················· ···················· ····················
Y 378 hCoV-19/Wales/PHWC-49786C/2020|EPI_ISL_726892|2020 ···················· ···················· ···················· ···················· ···················· ····················
Y 379 hCoV-19/Wales/PHWC-49787B/2020|EPI_ISL_726893|2020 ···················· ···················· ···················· ···················· ···················· ····················
Y 380 hCoV-19/Wales/PHWC-49788A/2020|EPI_ISL_726894|2020 ···················· ···················· ···················· ···················· ···················· ····················
Y 381 hCoV-19/Wales/PHWC-497899/2020|EPI_ISL_726895|2020 ···················· ···················· ···················· ···················· ···················· ····················
Y 382 hCoV-19/Wales/PHWC-4978A8/2020|EPI_ISL_726896|2020 ···················· ···················· ···················· ···················· ···················· ····················
Y 383 hCoV-19/Wales/PHWC-4978B7/2020|EPI_ISL_726897|2020 ···················· ···················· ···················· ···················· ···················· ····················
Y 384 hCoV-19/Wales/PHWC-4978C6/2020|EPI_ISL_726898|2020 ···················· ···················· ···················· ···················· ···················· ····················
Y 385 hCoV-19/Wales/PHWC-4978D5/2020|EPI_ISL_726899|2020 ···················· ···················· ···················· ···················· ···················· ····················
Y 386 hCoV-19/Switzerland/SG-ETHZ-390425/2020|EPI_ISL_72 ···················· ···················· ···················· ···················· ···················· ····················
Y 387 hCoV-19/Switzerland/BL-ETHZ-400394/2020|EPI_ISL_72 ···················· ···················· ···················· ···················· ···················· ····················
Y 388 hCoV-19/Switzerland/SO-ETHZ-400303/2020|EPI_ISL_72 ···················· ···················· ···················· ···················· ···················· ····················
Y 389 hCoV-19/Switzerland/BL-ETHZ-360269/2020|EPI_ISL_72 ···················· ···················· ···················· ···················· ···················· ····················
Y 390 hCoV-19/Switzerland/SH-ETHZ-400020/2020|EPI_ISL_72 ···················· ···················· ···················· ···················· ···················· ····················
Y 391 hCoV-19/Switzerland/BE-ETHZ-400386/2020|EPI_ISL_72 ···················· ···················· ···················· ···················· ···················· ····················
Y 392 hCoV-19/Switzerland/VD-ETHZ-360312/2020|EPI_ISL_72 ···················· ···················· ···················· ···················· ···················· ····················
Y 393 hCoV-19/Switzerland/BL-ETHZ-360249/2020|EPI_ISL_72 ···················· ···················· ···················· ···················· ···················· ····················
Y 394 hCoV-19/Switzerland/GR-ETHZ-400291/2020|EPI_ISL_72 ···················· ···················· ···················· ···················· ···················· ····················
Y 395 hCoV-19/Switzerland/GR-ETHZ-400292/2020|EPI_ISL_72 ···················· ···················· ···················· ···················· ···················· ····················
Y 396 hCoV-19/Switzerland/BE-ETHZ-390437/2020|EPI_ISL_72 ···················· ···················· ···················· ···················· ···················· ····················
Y 397 hCoV-19/Switzerland/BS-ETHZ-390438/2020|EPI_ISL_72 ···················· ···················· ···················· ···················· ···················· ····················
Y 398 hCoV-19/Switzerland/ZH-ETHZ-390439/2020|EPI_ISL_72 ···················· ···················· ···················· ···················· ···················· ····················
Y 399 hCoV-19/Switzerland/SZ-ETHZ-390440/2020|EPI_ISL_72 ···················· ···················· ···················· ···················· ···················· ····················
Y 400 hCoV-19/Switzerland/ZH-ETHZ-390441/2020|EPI_ISL_72 ···················· ···················· ···················· ···················· ···················· ····················
Y 401 hCoV-19/Switzerland/GR-ETHZ-390442/2020|EPI_ISL_72 ···················· ···················· ···················· ···················· ···················· ····················
Y 402 hCoV-19/Switzerland/SZ-ETHZ-390443/2020|EPI_ISL_72 ···················· ···················· ···················· ···················· ···················· ····················
Y 403 hCoV-19/Switzerland/SZ-ETHZ-390444/2020|EPI_ISL_72 ···················· ···················· ···················· ···················· ···················· ····················
Y 404 hCoV-19/Switzerland/AG-ETHZ-400395/2020|EPI_ISL_72 ···················· ···················· ···················· ···················· ···················· ····················
Y 405 hCoV-19/Switzerland/BL-ETHZ-400397/2020|EPI_ISL_72 ···················· ·A·················· ···················· ···················· ···················· ····················
Y 406 hCoV-19/Switzerland/SH-ETHZ-400021/2020|EPI_ISL_72 ···················· ·A·················· ···················· ···················· ···················· ····················
Y 407 hCoV-19/Switzerland/AG-ETHZ-400023/2020|EPI_ISL_72 ···················· ···················· ···················· ···················· ···················· ····················
Y 408 hCoV-19/Switzerland/ZH-ETHZ-400024/2020|EPI_ISL_72 ···················· ···················· ···················· ···················· ···················· ····················
Y 409 hCoV-19/Switzerland/TG-ETHZ-400025/2020|EPI_ISL_72 ···················· ···················· ···················· ···················· ···················· ····················
Y 410 hCoV-19/Switzerland/ZH-ETHZ-400028/2020|EPI_ISL_72 ···················· ···················· ···················· ···················· ···················· ····················
Y 411 hCoV-19/Switzerland/ZH-ETHZ-400029/2020|EPI_ISL_72 ···················· ···················· ···················· ···················· ···················· ····················
Y 412 hCoV-19/Switzerland/BE-ETHZ-400030/2020|EPI_ISL_72 ···················· ···················· ···················· ···················· ···················· ····················
Y 413 hCoV-19/Switzerland/BL-ETHZ-400031/2020|EPI_ISL_72 ···················· ···················· ···················· ···················· ···················· ····················
Y 414 hCoV-19/Switzerland/GR-ETHZ-400035/2020|EPI_ISL_72 ···················· ···················· ···················· ···················· ···················· ····················
Y 415 hCoV-19/Switzerland/BL-ETHZ-400036/2020|EPI_ISL_72 ···················· ···················· ···················· ···················· ···················· ····················
Y 416 hCoV-19/Nigeria/LA-CV367/2020|EPI_ISL_730028|2020- ···················· ···················· ···················· ···················· ···················· ····················
Y 417 hCoV-19/Nigeria/LA-CV368/2020|EPI_ISL_730029|2020- ···················· ···················· ···················· ···················· ···················· ····················
Y 418 hCoV-19/Nigeria/LA-CV370/2020|EPI_ISL_730031|2020- ···················· ···················· ···················· ···················· ···················· ····················
Y 419 hCoV-19/Nigeria/LA-CV371/2020|EPI_ISL_730032|2020- ···················· ···················· ···················· ···················· ···················· ····················
Y 420 hCoV-19/Nigeria/LA-CV374/2020|EPI_ISL_730033|2020- ···················· ···················· ···················· ···················· ···················· ····················
Y 421 hCoV-19/Nigeria/LA-CV375/2020|EPI_ISL_730034|2020- ···················· ···················· ···················· ···················· ···················· ····················
Y 422 hCoV-19/Nigeria/LA-CV379/2020|EPI_ISL_730035|2020- ···················· ···················· ···················· ···················· ···················· ····················
Y 423 hCoV-19/Nigeria/LA-CV380/2020|EPI_ISL_730036|2020- ···················· ···················· ···················· ···················· ···················· ····················
Y 424 hCoV-19/Nigeria/LA-CV384/2020|EPI_ISL_730038|2020- ···················· ···················· ···················· ···················· ···················· ····················
Y 425 hCoV-19/Nigeria/LA-CV386/2020|EPI_ISL_730039|2020- ···················· ···················· ···················· ···················· ···················· ····················
Y 426 hCoV-19/Nigeria/LA-CV389/2020|EPI_ISL_730042|2020- ···················· ···················· ···················· ···················· ···················· ····················
Y 427 hCoV-19/Nigeria/KW-CV394/2020|EPI_ISL_730044|2020- ···················· ···················· ···················· ···················· ···················· ····················
Y 428 hCoV-19/Nigeria/KW-CV401/2020|EPI_ISL_730048|2020- ···················· ···················· ···················· ···················· ···················· ····················
Y 429 hCoV-19/USA/CA-ALSR-4692/2020|EPI_ISL_730085|2020- ···················· ·A·················· ···················· ···················· ···················· ····················
Y 430 hCoV-19/USA/CA-ALSR-4693/2020|EPI_ISL_730086|2020- ···················· ···················· ···················· ···················· ···················· ····················
Y 431 hCoV-19/USA/CA-ALSR-4694/2020|EPI_ISL_730087|2020- ···················· ···················· ···················· ···················· ···················· ····················
Y 432 hCoV-19/USA/CA-ALSR-4699/2020|EPI_ISL_730088|2020- ···················· ·A·················· ···················· ···················· ···················· ····················
Y 433 hCoV-19/USA/CA-ALSR-4701/2020|EPI_ISL_730089|2020- ···················· ·A·················· ···················· ···················· ···················· ····················
Y 434 hCoV-19/USA/CA-ALSR-4702/2020|EPI_ISL_730090|2020- ···················· ·A·················· ···················· ···················· ···················· ····················
Y 435 hCoV-19/USA/CA-ALSR-4703/2020|EPI_ISL_730091|2020- ···················· ···················· ···················· ···················· ···················· ····················
Y 436 hCoV-19/USA/CA-ALSR-4704/2020|EPI_ISL_730092|2020- ···················· ···················· ···················· ···················· ···················· ····················
```

```
Y   437 hCoV-19/USA/CA-ALSR-4705/2020|EPI_ISL_730093|2020-   ····················   ·A··················   ····················   ····················   ····················   ····················
Y   438 hCoV-19/USA/CA-ALSR-4707/2020|EPI_ISL_730094|2020-   ····················   ····················   ····················   ····················   ····················   ····················
Y   439 hCoV-19/USA/CA-ALSR-4708/2020|EPI_ISL_730095|2020-   ····················   ·A··················   ····················   ····················   ····················   ····················
Y   440 hCoV-19/USA/CA-ALSR-4709/2020|EPI_ISL_730096|2020-   ····················   ·A··················   ····················   ····················   ····················   ····················
Y   441 hCoV-19/USA/CA-ALSR-4710/2020|EPI_ISL_730097|2020-   ····················   ·A··················   ····················   ····················   ····················   ····················
Y   442 hCoV-19/USA/CA-ALSR-4711/2020|EPI_ISL_730098|2020-   ····················   ····················   ····················   ····················   ····················   ····················
Y   443 hCoV-19/USA/CA-ALSR-4713/2020|EPI_ISL_730099|2020-   ····················   ····················   ····················   ····················   ····················   ····················
Y   444 hCoV-19/England/ALDP-C395F4/2020|EPI_ISL_731360|20   ····················   ····················   ····················   ····················   ····················   ····················
Y   445 hCoV-19/England/ALDP-C39390/2020|EPI_ISL_731363|20   ····················   ····················   ····················   ····················   ····················   ····················
Y   446 hCoV-19/England/ALDP-C3915A/2020|EPI_ISL_731364|20   ····················   ····················   ····················   ····················   ····················   ····················
Y   447 hCoV-19/England/ALDP-C39A5F/2020|EPI_ISL_731365|20   ····················   ····················   ····················   ····················   ····················   ····················
Y   448 hCoV-19/England/QEUH-C31EF1/2020|EPI_ISL_731367|20   ····················   ····················   ····················   ····················   ····················   ····················
Y   449 hCoV-19/England/QEUH-C315B0/2020|EPI_ISL_731368|20   ····················   ····················   ····················   ····················   ····················   ····················
Y   450 hCoV-19/England/ALDP-C3961F/2020|EPI_ISL_731369|20   ····················   ····················   ····················   ····················   ····················   ····················
Y   451 hCoV-19/England/ALDP-C394D9/2020|EPI_ISL_731371|20   ····················   ····················   ····················   ····················   ····················   ····················
Y   452 hCoV-19/England/ALDP-C3A766/2020|EPI_ISL_731375|20   ····················   ····················   ····················   ····················   ····················   ····················
Y   453 hCoV-19/England/QEUH-C312E6/2020|EPI_ISL_731378|20   ····················   ····················   ····················   ····················   ····················   ····················
Y   454 hCoV-19/England/QEUH-C30B91/2020|EPI_ISL_731379|20   ····················   ····················   ····················   ····················   ····················   ····················
Y   455 hCoV-19/England/QEUH-C31231/2020|EPI_ISL_731380|20   ····················   ····················   ····················   ····················   ····················   ····················
Y   456 hCoV-19/England/QEUH-C30AA3/2020|EPI_ISL_731383|20   ····················   ····················   ····················   ····················   ····················   ····················
Y   457 hCoV-19/France/ARA-111424/2020|EPI_ISL_732690|2020   ····················   ····················   ····················   ····················   ····················   ····················
Y   458 hCoV-19/France/ARA-111572/2020|EPI_ISL_732691|2020   ····················   ····················   ····················   ····················   ····················   ····················
Y   459 hCoV-19/France/ARA-111841/2020|EPI_ISL_732692|2020   ····················   ····················   ····················   ····················   ····················   ····················
Y   460 hCoV-19/France/ARA-75541/2020|EPI_ISL_732693|2020-   ····················   ····················   ····················   ····················   ····················   ····················
Y   461 hCoV-19/France/ARA-79153/2020|EPI_ISL_732694|2020-   ····················   ····················   ····················   ····················   ····················   ····················
Y   462 hCoV-19/France/ARA-83616/2020|EPI_ISL_732695|2020-   ····················   ····················   ····················   ····················   ····················   ····················
Y   463 hCoV-19/France/ARA-86532/2020|EPI_ISL_732696|2020-   ····················   ····················   ····················   ····················   ····················   ····················
Y   464 hCoV-19/France/ARA-86645/2020|EPI_ISL_732697|2020-   ····················   ····················   ····················   ····················   ····················   ····················
Y   465 hCoV-19/France/ARA-87533/2020|EPI_ISL_732698|2020-   ····················   ····················   ····················   ····················   ····················   ···············A····
Y   466 hCoV-19/France/ARA-95401/2020|EPI_ISL_732699|2020-   ····················   ····················   ····················   ····················   ····················   ····················
Y   467 hCoV-19/France/ARA-96089/2020|EPI_ISL_732701|2020-   ····················   ····················   ····················   ····················   ····················   ····················
Y   468 hCoV-19/France/ARA-96707/2020|EPI_ISL_732702|2020-   ····················   ····················   ····················   ····················   ····················   ····················
Y   469 hCoV-19/France/ARA-98766/2020|EPI_ISL_732703|2020-   ····················   ····················   ····················   ····················   ····················   ····················
Y   470 hCoV-19/USA/NM-DOH-2020314707/2020|EPI_ISL_732704|   ····················   ····················   ····················   ····················   ····················   ····················
Y   471 hCoV-19/USA/NM-DOH-2020315157/2020|EPI_ISL_732705|   ····················   ····················   ····················   ····················   ····················   ····················
Y   472 hCoV-19/USA/NM-DOH-2020315192/2020|EPI_ISL_732706|   ····················   ·A··················   ····················   ····················   ····················   ····················
Y   473 hCoV-19/USA/NM-DOH-2020315222/2020|EPI_ISL_732707|   ····················   ····················   ····················   ····················   ····················   ····················
Y   474 hCoV-19/USA/NM-DOH-2020315376/2020|EPI_ISL_732708|   ····················   ····················   ····················   ····················   ····················   ····················
Y   475 hCoV-19/USA/NM-DOH-2020315506/2020|EPI_ISL_732710|   ····················   ····················   ····················   ····················   ····················   ····················
Y   476 hCoV-19/USA/NM-DOH-2020315507/2020|EPI_ISL_732711|   ····················   ····················   ····················   ····················   ····················   ····················
Y   477 hCoV-19/USA/NM-DOH-2020315536/2020|EPI_ISL_732712|   ····················   ····················   ····················   ····················   ····················   ····················
Y   478 hCoV-19/USA/NM-DOH-2020315751/2020|EPI_ISL_732714|   ····················   ·A··················   ····················   ····················   ····················   ····················
Y   479 hCoV-19/USA/NM-DOH-2020315753/2020|EPI_ISL_732715|   ····················   ·A··················   ····················   ····················   ····················   ····················
Y   480 hCoV-19/USA/NM-DOH-2020315778/2020|EPI_ISL_732716|   ····················   ····················   ····················   ····················   ····················   ····················
Y   481 hCoV-19/USA/NM-DOH-2020316054/2020|EPI_ISL_732718|   ····················   ····················   ····················   ····················   ····················   ····················
Y   482 hCoV-19/USA/NM-DOH-2020316085/2020|EPI_ISL_732719|   ····················   ····················   ····················   ····················   ····················   ····················
Y   483 hCoV-19/USA/NM-DOH-2020316507/2020|EPI_ISL_732720|   ····················   ····················   ····················   ····················   ····················   ····················
Y   484 hCoV-19/USA/NM-DOH-2020316600/2020|EPI_ISL_732721|   ····················   ····················   ····················   ····················   ····················   ····················
Y   485 hCoV-19/USA/NM-DOH-2020316601/2020|EPI_ISL_732722|   ····················   ····················   ····················   ····················   ····················   ····················
Y   486 hCoV-19/England/QEUH-C57A5B/2020|EPI_ISL_736209|20   ····················   ····················   ····················   ····················   ····················   ····················
Y   487 hCoV-19/England/QEUH-C53FE7/2020|EPI_ISL_736212|20   ····················   ····················   ····················   ····················   ······T·············   ····················
Y   488 hCoV-19/England/MILK-C52C3C/2020|EPI_ISL_736219|20   ····················   ····················   ····················   ····················   ····················   ····················
Y   489 hCoV-19/England/QEUH-C57B2B/2020|EPI_ISL_736221|20   ····················   ····················   ····················   ····················   ····················   ····················
Y   490 hCoV-19/England/QEUH-C57B85/2020|EPI_ISL_736223|20   ····················   ····················   ····················   ····················   ····················   ····················
Y   491 hCoV-19/England/MILK-C52BE4/2020|EPI_ISL_736230|20   ····················   ····················   ····················   ····················   ····················   ····················
Y   492 hCoV-19/England/QEUH-C5406B/2020|EPI_ISL_736231|20   ····················   ····················   ····················   ····················   ····················   ····················
Y   493 hCoV-19/England/MILK-C52D0C/2020|EPI_ISL_736232|20   ····················   ····················   ····················   ····················   ····················   ····················
Y   494 hCoV-19/England/QEUH-C54247/2020|EPI_ISL_736235|20   ····················   ····················   ····················   ····················   ····················   ····················
Y   495 hCoV-19/England/QEUH-C57B67/2020|EPI_ISL_736236|20   ····················   ····················   ····················   ····················   ····················   ····················
Y   496 hCoV-19/England/QEUH-C544BA/2020|EPI_ISL_736238|20   ····················   ····················   ····················   ····················   ······T·············   ····················
Y   497 hCoV-19/England/QEUH-C5402F/2020|EPI_ISL_736242|20   ····················   ····················   ····················   ····················   ····················   ····················
Y   498 hCoV-19/England/QEUH-C57BC1/2020|EPI_ISL_736243|20   ····················   ····················   ····················   ····················   ······T·············   ····················
Y   499 hCoV-19/England/MILK-C52CE1/2020|EPI_ISL_736249|20   ····················   ····················   ····················   ····················   ····················   ····················
Y   500 hCoV-19/England/QEUH-C57AE2/2020|EPI_ISL_736250|20   ····················   ····················   ····················   ····················   ····················   ····················
Y   501 hCoV-19/England/MILK-C52C96/2020|EPI_ISL_736251|20   ····················   ····················   ····················   ····················   ····················   ····················
Y   502 hCoV-19/England/QEUH-C57B0D/2020|EPI_ISL_736255|20   ····················   ····················   ····················   ····················   ····················   ····················
Y   503 hCoV-19/England/QEUH-C546A5/2020|EPI_ISL_736257|20   ····················   ····················   ····················   ····················   ····················   ····················
Y   504 hCoV-19/England/QEUH-C57BFE/2020|EPI_ISL_736261|20   ····················   ····················   ····················   ····················   ····················   ····················
Y   505 hCoV-19/England/MILK-C52CF0/2020|EPI_ISL_736262|20   ····················   ····················   ····················   ····················   ····················   ····················
Y   506 hCoV-19/England/MILK-C52CB4/2020|EPI_ISL_736265|20   ····················   ····················   ····················   ····················   ····················   ····················
Y   507 hCoV-19/England/QEUH-C57A79/2020|EPI_ISL_736266|20   ····················   ····················   ····················   ····················   ····················   ····················
Y   508 hCoV-19/England/MILK-C525AA/2020|EPI_ISL_736267|20   ····················   ····················   ····················   ····················   ····················   ····················
Y   509 hCoV-19/England/QEUH-C542A1/2020|EPI_ISL_736275|20   ····················   ····················   ····················   ····················   ····················   ····················
Y   510 hCoV-19/England/QEUH-C53E80/2020|EPI_ISL_736277|20   ····················   ····················   ····················   ····················   ····················   ····················
Y   511 hCoV-19/England/MILK-C52D1B/2020|EPI_ISL_736279|20   ····················   ····················   ····················   ····················   ····················   ····················
Y   512 hCoV-19/England/MILK-C52A23/2020|EPI_ISL_736286|20   ····················   ····················   ····················   ····················   ····················   ····················
Y   513 hCoV-19/Wales/PHWC-49792D/2020|EPI_ISL_726904|2020   ····················   ····················   ····················   ····················   ····················   ····················
Y   515 hCoV-19/Wales/PHWC-49794B/2020|EPI_ISL_726906|2020   ····················   ····················   ····················   ····················   ····················   ····················
Y   516 hCoV-19/Wales/PHWC-49795A/2020|EPI_ISL_726907|2020   ····················   ····················   ····················   ····················   ····················   ····················
Y   517 hCoV-19/Wales/PHWC-497969/2020|EPI_ISL_726908|2020   ····················   ····················   ····················   ····················   ····················   ····················
```

| | | | | | | | |
|---|---|---|---|---|---|---|---|
| Y | 518 hCoV-19/Wales/PHWC-497978/2020\|EPI_ISL_726909\|2020 | ···················· | ···················· | ····················· | ···················· | ···················· | ···················· |
| Y | 519 hCoV-19/Wales/PHWC-497987/2020\|EPI_ISL_726910\|2020 | ···················· | ···················· | ····················· | ···················· | ···················· | ···················· |
| Y | 520 hCoV-19/Wales/PHWC-497996/2020\|EPI_ISL_726911\|2020 | ···················· | ···················· | ····················· | ···················· | ···················· | ···················· |
| Y | 521 hCoV-19/Wales/PHWC-4979A5/2020\|EPI_ISL_726912\|2020 | ···················· | ···················· | ····················· | ···················· | ···················· | ···················· |
| Y | 522 hCoV-19/Wales/PHWC-4979B4/2020\|EPI_ISL_726913\|2020 | ···················· | ···················· | ····················· | ···················· | ···················· | ···················· |
| Y | 524 hCoV-19/Wales/PHWC-497A2A/2020\|EPI_ISL_726920\|2020 | ···················· | ···················· | ····················· | ···················· | ···················· | ···················· |
| Y | 525 hCoV-19/Wales/PHWC-497A39/2020\|EPI_ISL_726921\|2020 | ···················· | ···················· | ····················· | ···················· | ···················· | ···················· |
| Y | 526 hCoV-19/Wales/PHWC-497A48/2020\|EPI_ISL_726922\|2020 | ···················· | ···················· | ····················· | ···················· | ···················· | ···················· |
| Y | 527 hCoV-19/Wales/PHWC-497A57/2020\|EPI_ISL_726923\|2020 | ···················· | ···················· | ····················· | ···················· | ···················· | ···················· |
| Y | 528 hCoV-19/Wales/PHWC-497A66/2020\|EPI_ISL_726924\|2020 | ···················· | ···················· | ····················· | ···················· | ···················· | ···················· |
| Y | 529 hCoV-19/Wales/PHWC-497A75/2020\|EPI_ISL_726925\|2020 | ···················· | ···················· | ····················· | ···················· | ···················· | ···················· |
| Y | 530 hCoV-19/Switzerland/SO-ETHZ-400040/2020\|EPI_ISL_72 | ···················· | ···················· | ····················· | ···················· | ···················· | ···K················ |
| Y | 532 hCoV-19/Switzerland/SO-ETHZ-400042/2020\|EPI_ISL_72 | ···················· | ···················· | ····················· | ···················· | ···················· | ···················· |
| Y | 533 hCoV-19/Switzerland/SO-ETHZ-400043/2020\|EPI_ISL_72 | ···················· | ···················· | ····················· | ···················· | ···················· | ···················· |
| Y | 534 hCoV-19/Switzerland/AG-ETHZ-400045/2020\|EPI_ISL_72 | ···················· | ···················· | ····················· | ···················· | ···················· | ···················· |
| Y | 535 hCoV-19/Switzerland/SO-ETHZ-400046/2020\|EPI_ISL_72 | ···················· | ···················· | ····················· | ···················· | ······K············· | ···················· |
| Y | 536 hCoV-19/Switzerland/SO-ETHZ-400047/2020\|EPI_ISL_72 | ···················· | ···················· | ····················· | ···················· | ···················· | ···················· |
| Y | 537 hCoV-19/Switzerland/SO-ETHZ-400048/2020\|EPI_ISL_72 | ···················· | ···················· | ····················· | ···················· | ···················· | ···················· |
| Y | 538 hCoV-19/Switzerland/SO-ETHZ-400050/2020\|EPI_ISL_72 | ···················· | ···················· | ····················· | ···················· | ···················· | ···················· |
| Y | 539 hCoV-19/Switzerland/ZH-ETHZ-400052/2020\|EPI_ISL_72 | ···················· | ···················· | ····················· | ···················· | ···S················ | ···················· |
| Y | 540 hCoV-19/Switzerland/BE-ETHZ-400053/2020\|EPI_ISL_72 | ···················· | ···················· | ····················· | ···················· | ···················· | ···················· |
| Y | 541 hCoV-19/Switzerland/BE-ETHZ-400055/2020\|EPI_ISL_72 | ···················· | ···················· | ····················· | ···················· | ···················· | ···················· |
| Y | 542 hCoV-19/Switzerland/BE-ETHZ-400056/2020\|EPI_ISL_72 | ···················· | ···················· | ····················· | ···················· | ···················· | ···················· |
| Y | 543 hCoV-19/Switzerland/ETHZ-400057/2020\|EPI_ISL_72911 | ···················· | ···················· | ····················· | ···················· | ···················· | ···················· |
| Y | 544 hCoV-19/Switzerland/BE-ETHZ-400060/2020\|EPI_ISL_72 | ···················· | ···················· | ····················· | ···················· | ···················· | ···················· |
| Y | 545 hCoV-19/Switzerland/BE-ETHZ-400062/2020\|EPI_ISL_72 | ···················· | ···················· | ····················· | ···················· | ···················· | ···················· |
| Y | 546 hCoV-19/Switzerland/BE-ETHZ-400063/2020\|EPI_ISL_72 | ···················· | ···················· | ····················· | ···················· | ···················· | ···················· |
| Y | 547 hCoV-19/Switzerland/BE-ETHZ-400064/2020\|EPI_ISL_72 | ···················· | ···················· | ····················· | ···················· | ···················· | ···················· |
| Y | 548 hCoV-19/Switzerland/BE-ETHZ-400066/2020\|EPI_ISL_72 | ···················· | ···················· | ····················· | ···················· | ···················· | ···················· |
| Y | 549 hCoV-19/Switzerland/BE-ETHZ-400067/2020\|EPI_ISL_72 | ···················· | ···················· | ····················· | ···················· | ···················· | ···················· |
| Y | 550 hCoV-19/Switzerland/BE-ETHZ-400068/2020\|EPI_ISL_72 | ···················· | ······A············· | ····················· | ···················· | ···················· | ···R················ |
| Y | 551 hCoV-19/Switzerland/ZH-ETHZ-400069/2020\|EPI_ISL_72 | ···················· | ···················· | ····················· | ···················· | ···················· | ···················· |
| Y | 552 hCoV-19/Switzerland/BE-ETHZ-400070/2020\|EPI_ISL_72 | ···················· | ···················· | ····················· | ···················· | ···················· | ···················· |
| Y | 553 hCoV-19/Switzerland/BE-ETHZ-400071/2020\|EPI_ISL_72 | ···················· | ···················· | ····················· | ···················· | ······K············· | ···················· |
| Y | 554 hCoV-19/Switzerland/BE-ETHZ-400073/2020\|EPI_ISL_72 | ···················· | ···················· | ····················· | ···················· | ···················· | ···················· |
| Y | 555 hCoV-19/Switzerland/BE-ETHZ-400074/2020\|EPI_ISL_72 | ···················· | ···················· | ····················· | ···················· | ···················· | ···················· |
| Y | 556 hCoV-19/Switzerland/BE-ETHZ-400075/2020\|EPI_ISL_72 | ···················· | ·A·················· | ····················· | ···················· | ···················· | ···A················ |
| Y | 557 hCoV-19/Switzerland/BE-ETHZ-400078/2020\|EPI_ISL_72 | ···················· | ···················· | ····················· | ···················· | ···················· | ···················· |
| Y | 558 hCoV-19/Switzerland/BE-ETHZ-400079/2020\|EPI_ISL_72 | ···················· | ···················· | ····················· | ···················· | ···················· | ···················· |
| Y | 559 hCoV-19/Switzerland/BE-ETHZ-400080/2020\|EPI_ISL_72 | ···················· | ···················· | ····················· | ···················· | ···················· | ···················· |
| Y | 560 hCoV-19/Switzerland/BS-ETHZ-400081/2020\|EPI_ISL_72 | ···················· | ·A·················· | ····················· | ···················· | ···················· | ···R················ |
| Y | 561 hCoV-19/Switzerland/BE-ETHZ-400082/2020\|EPI_ISL_72 | ···················· | ···················· | ····················· | ···················· | ···················· | ···················· |
| Y | 562 hCoV-19/Switzerland/BE-ETHZ-400083/2020\|EPI_ISL_72 | ···················· | ···················· | ····················· | ···················· | ···················· | ···················· |
| Y | 563 hCoV-19/Switzerland/BE-ETHZ-400085/2020\|EPI_ISL_72 | ···················· | ···················· | ····················· | ···················· | ···················· | ···················· |
| Y | 564 hCoV-19/Switzerland/BE-ETHZ-400086/2020\|EPI_ISL_72 | ···················· | ···················· | ····················· | ···················· | ···················· | ···················· |
| Y | 565 hCoV-19/Switzerland/BE-ETHZ-400087/2020\|EPI_ISL_72 | ···················· | ···················· | ····················· | ···················· | ···················· | ···················· |
| Y | 566 hCoV-19/Switzerland/BE-ETHZ-400089/2020\|EPI_ISL_72 | ···················· | ···················· | ····················· | ···················· | ···················· | ···················· |
| Y | 567 hCoV-19/Switzerland/BE-ETHZ-400091/2020\|EPI_ISL_72 | ···················· | ···················· | ····················· | ···················· | ···················· | ···················· |
| Y | 568 hCoV-19/Switzerland/BL-ETHZ-400092/2020\|EPI_ISL_72 | ···················· | ···················· | ····················· | ···················· | ···················· | ···················· |
| Y | 569 hCoV-19/Switzerland/BL-ETHZ-400094/2020\|EPI_ISL_72 | ···················· | ···················· | ····················· | ···················· | ···················· | ···················· |
| Y | 570 hCoV-19/Switzerland/BL-ETHZ-400095/2020\|EPI_ISL_72 | ···················· | ···················· | ····················· | ···················· | ···················· | ···················· |
| Y | 571 hCoV-19/Switzerland/BL-ETHZ-400096/2020\|EPI_ISL_72 | ···················· | ···················· | ····················· | ···················· | ···················· | ···················· |
| Y | 572 hCoV-19/Switzerland/SO-ETHZ-400098/2020\|EPI_ISL_72 | ···················· | ···················· | ····················· | ···················· | ···················· | ···················· |
| Y | 573 hCoV-19/Switzerland/BL-ETHZ-400101/2020\|EPI_ISL_72 | ···················· | ···················· | ····················· | ···················· | ···················· | ···················· |
| Y | 574 hCoV-19/Switzerland/BL-ETHZ-400102/2020\|EPI_ISL_72 | ···················· | ···················· | ····················· | ···················· | ···················· | ···················· |
| Y | 575 hCoV-19/Switzerland/BE-ETHZ-400103/2020\|EPI_ISL_72 | ···················· | ···················· | ····················· | ···················· | ···················· | ···················· |
| Y | 576 hCoV-19/Switzerland/BL-ETHZ-400104/2020\|EPI_ISL_72 | ···················· | ···················· | ····················· | ···················· | ···················· | ···················· |
| Y | 577 hCoV-19/Switzerland/BE-ETHZ-400105/2020\|EPI_ISL_72 | ···················· | ···················· | ····················· | ···················· | ···················· | ···················· |
| Y | 578 hCoV-19/Switzerland/SO-ETHZ-400106/2020\|EPI_ISL_72 | ···················· | ···················· | ····················· | ···················· | ···················· | ···················· |
| Y | 579 hCoV-19/Switzerland/BL-ETHZ-400107/2020\|EPI_ISL_72 | ···················· | ···················· | ····················· | ···················· | ···················· | ···················· |
| Y | 580 hCoV-19/Switzerland/BL-ETHZ-400108/2020\|EPI_ISL_72 | ···················· | ···················· | ····················· | ···················· | ···················· | ···················· |
| Y | 581 hCoV-19/Switzerland/ZH-ETHZ-400133/2020\|EPI_ISL_72 | ···················· | ···················· | ····················· | ···················· | ···················· | ···················· |
| Y | 582 hCoV-19/Switzerland/GR-ETHZ-400286/2020\|EPI_ISL_72 | ···················· | ···················· | ····················· | ···················· | ···················· | ···················· |
| Y | 583 hCoV-19/Switzerland/GR-ETHZ-400288/2020\|EPI_ISL_72 | ···················· | ···················· | ····················· | ···················· | ···················· | ···················· |
| Y | 584 hCoV-19/Switzerland/GR-ETHZ-400289/2020\|EPI_ISL_72 | ···················· | ···················· | ····················· | ···················· | ···················· | ···················· |
| Y | 585 hCoV-19/Switzerland/GR-ETHZ-400290/2020\|EPI_ISL_72 | ···················· | ······A············· | ····················· | ···················· | ···················· | ···················· |
| Y | 586 hCoV-19/Switzerland/SO-ETHZ-400297/2020\|EPI_ISL_72 | ···················· | ···················· | ····················· | ···················· | ···················· | ···················· |
| Y | 587 hCoV-19/Switzerland/SO-ETHZ-400299/2020\|EPI_ISL_72 | ···················· | ···················· | ····················· | ···················· | ···················· | ···················· |
| Y | 588 hCoV-19/Switzerland/SO-ETHZ-400300/2020\|EPI_ISL_72 | ···················· | ···················· | ····················· | ···················· | ···················· | ···················· |
| Y | 589 hCoV-19/Switzerland/SO-ETHZ-400301/2020\|EPI_ISL_72 | ···················· | ···················· | ····················· | ···················· | ···················· | ···················· |
| Y | 590 hCoV-19/Switzerland/BL-ETHZ-400370/2020\|EPI_ISL_72 | ···················· | ···················· | ····················· | ···················· | ···················· | ···················· |
| Y | 591 hCoV-19/Switzerland/AG-ETHZ-400371/2020\|EPI_ISL_72 | ···················· | ···················· | ····················· | ···················· | ···················· | ···················· |
| Y | 592 hCoV-19/Switzerland/LU-ETHZ-400372/2020\|EPI_ISL_72 | ···················· | ···················· | ····················· | ···················· | ······K············· | ···················· |
| Y | 593 hCoV-19/Switzerland/ZH-ETHZ-400373/2020\|EPI_ISL_72 | ···················· | ···················· | ····················· | ···················· | ···················· | ···················· |
| Y | 594 hCoV-19/Switzerland/LU-ETHZ-400375/2020\|EPI_ISL_72 | ···················· | ···················· | ····················· | ···················· | ······K············· | ···················· |
| Y | 595 hCoV-19/Switzerland/ZH-ETHZ-400377/2020\|EPI_ISL_72 | ···················· | ···················· | ····················· | ···················· | ···················· | ···················· |
| Y | 596 hCoV-19/Switzerland/BE-ETHZ-400382/2020\|EPI_ISL_72 | ···················· | ···················· | ····················· | ···················· | ···················· | ···················· |
| Y | 597 hCoV-19/Switzerland/BL-ETHZ-350293/2020\|EPI_ISL_72 | ···················· | ···················· | ····················· | ···················· | ···················· | ···················· |
| Y | 598 hCoV-19/Switzerland/AG-ETHZ-350309/2020\|EPI_ISL_72 | ···················· | ···················· | ····················· | ···················· | ···················· | ···R················ |
| Y | 599 hCoV-19/Switzerland/VS-ETHZ-350316/2020\|EPI_ISL_72 | ···················· | ······A············· | ····················· | ···················· | ···················· | ···················· |

| | | | | | | | | |
|---|---|---|---|---|---|---|---|---|
| Y | 600 | hCoV-19/Switzerland/NE-ETHZ-350323/2020\|EPI_ISL_72 | ···················· | ···················· | ····················· | ···················· | ···················· | ···················· |
| Y | 601 | hCoV-19/Switzerland/NE-ETHZ-350329/2020\|EPI_ISL_72 | ···················· | ···················· | ····················· | ···················· | ···················· | ···················· |
| Y | 602 | hCoV-19/Switzerland/VD-ETHZ-350330/2020\|EPI_ISL_72 | ···················· | ···················· | ····················· | ···················· | ···················· | ···················· |
| Y | 603 | hCoV-19/Switzerland/BE-ETHZ-350336/2020\|EPI_ISL_72 | ·······T············ | ···················· | ····················· | ···················· | ···················· | ···················· |
| Y | 604 | hCoV-19/Switzerland/BL-ETHZ-350342/2020\|EPI_ISL_72 | ···················· | ···················· | ····················· | ···················· | ···················· | ······R············· |
| Y | 605 | hCoV-19/Switzerland/AG-ETHZ-350351/2020\|EPI_ISL_72 | ···················· | ···················· | ····················· | ···················· | ···················· | ···················· |
| Y | 606 | hCoV-19/Switzerland/AG-ETHZ-350354/2020\|EPI_ISL_72 | ···········T········ | ···················· | ····················· | ···················· | ···················· | ···················· |
| Y | 607 | hCoV-19/USA/CA-ALSR-4715/2020\|EPI_ISL_730100\|2020- | ···················· | ···················· | ····················· | ···················· | ···················· | ···················· |
| Y | 608 | hCoV-19/USA/CA-ALSR-4716/2020\|EPI_ISL_730101\|2020- | ···················· | ·A·················· | ····················· | ···················· | ···················· | ···················· |
| Y | 609 | hCoV-19/USA/CA-ALSR-4718/2020\|EPI_ISL_730102\|2020- | ···················· | ·A·················· | ····················· | ···················· | ···················· | ···················· |
| Y | 610 | hCoV-19/USA/CA-ALSR-4719/2020\|EPI_ISL_730103\|2020- | ···················· | ·A·················· | ····················· | ···················· | ···················· | ···················· |
| Y | 611 | hCoV-19/USA/CA-ALSR-4721/2020\|EPI_ISL_730104\|2020- | ···················· | ···················· | ····················· | ···················· | ···················· | ···················· |
| Y | 612 | hCoV-19/USA/CA-ALSR-4722/2020\|EPI_ISL_730105\|2020- | ···················· | ·A·················· | ····················· | ···················· | ···················· | ···················· |
| Y | 613 | hCoV-19/USA/CA-ALSR-4723/2020\|EPI_ISL_730106\|2020- | ···················· | ·A·················· | ····················· | ···················· | ···················· | ···················· |
| Y | 614 | hCoV-19/USA/CA-ALSR-4724/2020\|EPI_ISL_730107\|2020- | ···················· | ···················· | ····················· | ···················· | ···················· | ···················· |
| Y | 615 | hCoV-19/USA/CA-ALSR-4725/2020\|EPI_ISL_730108\|2020- | ···················· | ···················· | ····················· | ···················· | ···················· | ···················· |
| Y | 616 | hCoV-19/USA/CA-ALSR-4726/2020\|EPI_ISL_730109\|2020- | ···················· | ···················· | ····················· | ···················· | ···················· | ···················· |
| Y | 617 | hCoV-19/USA/CA-ALSR-4727/2020\|EPI_ISL_730110\|2020- | ···················· | ·A·················· | ····················· | ···················· | ···················· | ···················· |
| Y | 618 | hCoV-19/USA/CA-ALSR-4728/2020\|EPI_ISL_730111\|2020- | ···················· | ···················· | ····················· | ···················· | ···················· | ···················· |
| Y | 619 | hCoV-19/USA/CA-ALSR-4730/2020\|EPI_ISL_730112\|2020- | ···················· | ·A·················· | ····················· | ···················· | ···················· | ···················· |
| Y | 620 | hCoV-19/USA/CA-ALSR-4734/2020\|EPI_ISL_730113\|2020- | ···················· | ·A·················· | ····················· | ···················· | ···················· | ···················· |
| Y | 621 | hCoV-19/USA/CA-ALSR-4735/2020\|EPI_ISL_730114\|2020- | ···················· | ·A·················· | ····················· | ···················· | ···················· | ···················· |
| Y | 622 | hCoV-19/USA/CA-ALSR-4736/2020\|EPI_ISL_730115\|2020- | ·······T············ | ·A·················· | ····················· | ···················· | ···················· | ···················· |
| Y | 623 | hCoV-19/USA/CA-ALSR-4738/2020\|EPI_ISL_730116\|2020- | ·······T············ | ·A·················· | ····················· | ···················· | ···················· | ···················· |
| Y | 624 | hCoV-19/USA/CA-ALSR-4739/2020\|EPI_ISL_730117\|2020- | ···················· | ···················· | ····················· | ···················· | ···················· | ···················· |
| Y | 625 | hCoV-19/USA/CA-ALSR-4744/2020\|EPI_ISL_730118\|2020- | ···················· | ·A·················· | ····················· | ···················· | ···················· | ···················· |
| Y | 626 | hCoV-19/USA/CA-ALSR-4749/2020\|EPI_ISL_730120\|2020- | ···················· | ·A·················· | ····················· | ···················· | ···················· | ···················· |
| Y | 627 | hCoV-19/USA/CA-ALSR-4750/2020\|EPI_ISL_730121\|2020- | ···················· | ···················· | ····················· | ···················· | ···················· | ···················· |
| Y | 628 | hCoV-19/USA/CA-ALSR-4751/2020\|EPI_ISL_730122\|2020- | ···················· | ···················· | ····················· | ···················· | ···················· | ···················· |
| Y | 629 | hCoV-19/USA/CA-ALSR-4754/2020\|EPI_ISL_730123\|2020- | ···················· | ·A·················· | ················G···· | ···················· | ···················· | ···················· |
| Y | 630 | hCoV-19/USA/CA-ALSR-4757/2020\|EPI_ISL_730124\|2020- | ···················· | ···················· | ····················· | ···················· | ···················· | ···················· |
| Y | 631 | hCoV-19/USA/CA-ALSR-4758/2020\|EPI_ISL_730125\|2020- | ···················· | ·A·················· | ····················· | ···················· | ···················· | ···················· |
| Y | 632 | hCoV-19/Mexico/BCN-ALSR-4762/2020\|EPI_ISL_730127\|2 | ···················· | ·A·················· | ····················· | ···················· | ···················· | ···················· |
| Y | 633 | hCoV-19/Mexico/BCN-ALSR-4766/2020\|EPI_ISL_730128\|2 | ···················· | ·A·················· | ····················· | ···················· | ···················· | ···················· |
| Y | 634 | hCoV-19/Mexico/BCN-ALSR-4768/2020\|EPI_ISL_730129\|2 | ···················· | ·A·················· | ····················· | ···················· | ···················· | ···················· |
| Y | 635 | hCoV-19/Mexico/BCN-ALSR-4770/2020\|EPI_ISL_730130\|2 | ···················· | ·A·················· | ····················· | ···················· | ···················· | ···················· |
| Y | 636 | hCoV-19/Mexico/BCN-ALSR-4772/2020\|EPI_ISL_730132\|2 | ···················· | ···················· | ····················· | ···················· | ···················· | ···················· |
| Y | 637 | hCoV-19/Mexico/BCN-ALSR-4774/2020\|EPI_ISL_730133\|2 | ···················· | ·A·················· | ····················· | ···················· | ···················· | ···················· |
| Y | 638 | hCoV-19/Mexico/BCN-ALSR-4775/2020\|EPI_ISL_730134\|2 | ···················· | ···················· | ····················· | ···················· | ···················· | ···················· |
| Y | 639 | hCoV-19/Mexico/BCN-ALSR-4778/2020\|EPI_ISL_730136\|2 | ···················· | ·A·················· | ····················· | ···················· | ···················· | ···················· |
| Y | 640 | hCoV-19/USA/CA-ALSR-4782/2020\|EPI_ISL_730138\|2020- | ···················· | ···················· | ····················· | ···················· | ···················· | ···················· |
| Y | 641 | hCoV-19/USA/CA-ALSR-4783/2020\|EPI_ISL_730139\|2020- | ···················· | ·A·················· | ····················· | ···················· | ···················· | ···················· |
| Y | 642 | hCoV-19/USA/CA-ALSR-4787/2020\|EPI_ISL_730140\|2020- | ···················· | ·A·················· | ····················· | ···················· | ···················· | ···················· |
| Y | 643 | hCoV-19/USA/CA-ALSR-4789/2020\|EPI_ISL_730141\|2020- | ···················· | ···················· | ····················· | ···················· | ···················· | ···················· |
| Y | 644 | hCoV-19/USA/CA-ALSR-4790/2020\|EPI_ISL_730142\|2020- | ···················· | ·A·················· | ····················· | ···················· | ···················· | ···················· |
| Y | 645 | hCoV-19/USA/CA-ALSR-4791/2020\|EPI_ISL_730143\|2020- | ···················· | ·A·················· | ····················· | ···················· | ···················· | ···················· |
| Y | 646 | hCoV-19/USA/CA-ALSR-4792/2020\|EPI_ISL_730144\|2020- | ···················· | ·A·················· | ····················· | ···················· | ···················· | ···················· |
| Y | 647 | hCoV-19/USA/CA-ALSR-4795/2020\|EPI_ISL_730146\|2020- | ···················· | ·A·················· | ····················· | ···················· | ···················· | ···················· |
| Y | 648 | hCoV-19/USA/CA-ALSR-4857/2020\|EPI_ISL_730180\|2020- | ···················· | ···················· | ····················· | ···················· | ···················· | ···················· |
| Y | 649 | hCoV-19/USA/CA-ALSR-4861/2020\|EPI_ISL_730181\|2020- | ···················· | ·A·················· | ····················· | ···················· | ···················· | ···················· |
| Y | 650 | hCoV-19/USA/CA-ALSR-4862/2020\|EPI_ISL_730182\|2020- | ···················· | ···················· | ····················· | ···················· | ···················· | ···················· |
| Y | 651 | hCoV-19/USA/CA-ALSR-4863/2020\|EPI_ISL_730183\|2020- | ···················· | ···················· | ····················· | ···················· | ···················· | ···················· |
| Y | 652 | hCoV-19/USA/CA-ALSR-4864/2020\|EPI_ISL_730184\|2020- | ···················· | ·A·················· | ····················· | ···················· | ···················· | ···················· |
| Y | 653 | hCoV-19/USA/CA-ALSR-4866/2020\|EPI_ISL_730186\|2020- | ···················· | ·A·················· | ····················· | ···················· | ···················· | ···················· |
| Y | 654 | hCoV-19/USA/CA-ALSR-4867/2020\|EPI_ISL_730187\|2020- | ···················· | ···················· | ····················· | ···················· | ···················· | ···················· |
| Y | 655 | hCoV-19/USA/CA-ALSR-4868/2020\|EPI_ISL_730188\|2020- | ···················· | ···················· | ····················· | ···················· | ···················· | ···················· |
| Y | 656 | hCoV-19/USA/CA-ALSR-4870/2020\|EPI_ISL_730189\|2020- | ···················· | ···················· | ····················· | ···················· | ···················· | ···················· |
| Y | 657 | hCoV-19/England/QEUH-C312F5/2020\|EPI_ISL_731400\|20 | ···················· | ···················· | ····················· | ···················· | ···················· | ···················· |
| Y | 658 | hCoV-19/England/QEUH-C30A58/2020\|EPI_ISL_731402\|20 | ···················· | ···················· | ····················· | ···················· | ······T············· | ···················· |
| Y | 659 | hCoV-19/England/QEUH-C3133E/2020\|EPI_ISL_731403\|20 | ···················· | ···················· | ····················· | ···················· | ···················· | ···················· |
| Y | 660 | hCoV-19/England/QEUH-C35F54/2020\|EPI_ISL_731404\|20 | ···················· | ···················· | ····················· | ···················· | ···················· | ···················· |
| Y | 661 | hCoV-19/England/QEUH-C311E9/2020\|EPI_ISL_731406\|20 | ···················· | ···················· | ····················· | ···················· | ···················· | ···················· |
| Y | 662 | hCoV-19/England/QEUH-C313E3/2020\|EPI_ISL_731411\|20 | ···················· | ···················· | ····················· | ···················· | ···················· | ···················· |
| Y | 663 | hCoV-19/England/QEUH-C3140E/2020\|EPI_ISL_731414\|20 | ···················· | ···················· | ····················· | ···················· | ···················· | ···················· |
| Y | 664 | hCoV-19/England/ALDP-C395C7/2020\|EPI_ISL_731415\|20 | ···················· | ···················· | ····················· | ···················· | ···················· | ···················· |
| Y | 665 | hCoV-19/England/ALDP-C3A511/2020\|EPI_ISL_731416\|20 | ···················· | ···················· | ····················· | ···················· | ···················· | ···················· |
| Y | 666 | hCoV-19/England/QEUH-C310DD/2020\|EPI_ISL_731417\|20 | ···················· | ···················· | ····················· | ···················· | ···················· | ···················· |
| Y | 667 | hCoV-19/England/ALDP-C3AB97/2020\|EPI_ISL_731418\|20 | ···················· | ···················· | ····················· | ···················· | ···················· | ···················· |
| Y | 668 | hCoV-19/England/ALDP-C39266/2020\|EPI_ISL_731419\|20 | ···················· | ···················· | ····················· | ···················· | ···················· | ···················· |
| Y | 669 | hCoV-19/England/QEUH-C35FEB/2020\|EPI_ISL_731420\|20 | ···················· | ···················· | ····················· | ···················· | ···················· | ···················· |
| Y | 670 | hCoV-19/England/QEUH-C30A85/2020\|EPI_ISL_731421\|20 | ···················· | ···················· | ····················· | ···················· | ···················· | ···················· |
| Y | 671 | hCoV-19/England/QEUH-C3151A/2020\|EPI_ISL_731426\|20 | ···················· | ···················· | ····················· | ···················· | ···················· | ···················· |
| Y | 672 | hCoV-19/England/ALDP-C3A748/2020\|EPI_ISL_731427\|20 | ···················· | ···················· | ····················· | ···················· | ···················· | ···················· |
| Y | 673 | hCoV-19/England/ALDP-C3A757/2020\|EPI_ISL_731428\|20 | ···················· | ···················· | ····················· | ···················· | ···················· | ···················· |
| Y | 674 | hCoV-19/England/QEUH-C31E79/2020\|EPI_ISL_731431\|20 | ···················· | ···················· | ····················· | ···················· | ···················· | ···················· |
| Y | 675 | hCoV-19/England/QEUH-C31741/2020\|EPI_ISL_731432\|20 | ···················· | ···················· | ····················· | ···················· | ···················· | ···················· |
| Y | 676 | hCoV-19/England/ALDP-C3AB2E/2020\|EPI_ISL_731434\|20 | ···················· | ···················· | ····················· | ···················· | ···················· | ···················· |
| Y | 677 | hCoV-19/England/QEUH-C31705/2020\|EPI_ISL_731435\|20 | ···················· | ···················· | ····················· | ···················· | ···················· | ···················· |
| Y | 678 | hCoV-19/England/ALDP-C395D6/2020\|EPI_ISL_731439\|20 | ···················· | ···················· | ····················· | ···················· | ···················· | ···················· |
| Y | 679 | hCoV-19/England/ALDP-C3A6A5/2020\|EPI_ISL_731440\|20 | ···················· | ···················· | ····················· | ···················· | ···················· | ···················· |

| | | | | | | | |
|---|---|---|---|---|---|---|---|
| Y | 680 hCoV-19/England/ALDP-C3A1B3/2020\|EPI_ISL_731441\|20 | ···················· | ···················· | ····················· | ···················· | ···················· | ···················· |
| Y | 681 hCoV-19/England/ALDP-C3A6D2/2020\|EPI_ISL_731442\|20 | ···················· | ···················· | ····················· | ···················· | ···················· | ···················· |
| Y | 682 hCoV-19/England/QEUH-C31161/2020\|EPI_ISL_731444\|20 | ···················· | ···················· | ····················· | ···················· | ···················· | ···················· |
| Y | 683 hCoV-19/England/ALDP-C3AD82/2020\|EPI_ISL_731446\|20 | ···················· | ···················· | ····················· | ···················· | ···················· | ···················· |
| Y | 684 hCoV-19/England/QEUH-C30B55/2020\|EPI_ISL_731447\|20 | ···················· | ···················· | ····················· | ···················· | ···················· | ···················· |
| Y | 685 hCoV-19/England/ALDP-C3A46F/2020\|EPI_ISL_731448\|20 | ···················· | ···················· | ····················· | ···················· | ···················· | ···················· |
| Y | 686 hCoV-19/England/QEUH-C31556/2020\|EPI_ISL_731449\|20 | ···················· | ···················· | ····················· | ···················· | ···················· | ···················· |
| Y | 687 hCoV-19/England/ALDP-C39503/2020\|EPI_ISL_731451\|20 | ···················· | ···················· | ····················· | ···················· | ···················· | ···················· |
| Y | 688 hCoV-19/England/QEUH-C30B37/2020\|EPI_ISL_731452\|20 | ···················· | ···················· | ····················· | ···················· | ···················· | ···················· |
| Y | 689 hCoV-19/England/QEUH-C316BD/2020\|EPI_ISL_731453\|20 | ···················· | ···················· | ····················· | ···················· | ···················· | ···················· |
| Y | 690 hCoV-19/England/ALDP-C3A793/2020\|EPI_ISL_731457\|20 | ···················· | ···················· | ····················· | ···················· | ···················· | ···················· |
| Y | 691 hCoV-19/England/QEUH-C35F81/2020\|EPI_ISL_731458\|20 | ···················· | ···················· | ····················· | ···················· | ···················· | ···················· |
| Y | 692 hCoV-19/England/QEUH-C30BBF/2020\|EPI_ISL_731459\|20 | ···················· | ···················· | ····················· | ···················· | ···················· | ···················· |
| Y | 693 hCoV-19/England/ALDP-C39309/2020\|EPI_ISL_731460\|20 | ···················· | ···················· | ··T·················· | ···················· | ···················· | ···················· |
| Y | 694 hCoV-19/England/ALDP-C3A61E/2020\|EPI_ISL_731461\|20 | ···················· | ···················· | ····················· | ···················· | ·····T·············· | ···················· |
| Y | 695 hCoV-19/England/ALDP-C3A423/2020\|EPI_ISL_731462\|20 | ···················· | ···················· | ····················· | ···················· | ···················· | ···················· |
| Y | 696 hCoV-19/England/ALDP-C3A687/2020\|EPI_ISL_731464\|20 | ···················· | ···················· | ····················· | ···················· | ···················· | ···················· |
| Y | 697 hCoV-19/England/ALDP-C3AD73/2020\|EPI_ISL_731465\|20 | ···················· | ···················· | ····················· | ···················· | ···················· | ···················· |
| Y | 698 hCoV-19/England/QEUH-C31240/2020\|EPI_ISL_731466\|20 | ···················· | ···················· | ····················· | ···················· | ···················· | ···················· |
| Y | 699 hCoV-19/England/ALDP-C3A1A4/2020\|EPI_ISL_731470\|20 | ···················· | ···················· | ····················· | ···················· | ···················· | ···················· |
| Y | 700 hCoV-19/England/ALDP-C392A2/2020\|EPI_ISL_731473\|20 | ···················· | ···················· | ····················· | ···················· | ···················· | ···················· |
| Y | 701 hCoV-19/England/QEUH-C30B19/2020\|EPI_ISL_731477\|20 | ···················· | ···················· | ····················· | ···················· | ···················· | ···················· |
| Y | 702 hCoV-19/England/QEUH-C30A49/2020\|EPI_ISL_731479\|20 | ···················· | ···················· | ····················· | ···················· | ···················· | ···················· |
| Y | 703 hCoV-19/France/ARA-96015/2020\|EPI_ISL_732700\|2020- | ···················· | ···················· | ····················· | ···················· | ···················· | ···················· |
| Y | 704 hCoV-19/USA/NM-DOH-2020316610/2020\|EPI_ISL_732723\| | ···················· | ···················· | ····················· | ···················· | ···················· | ···················· |
| Y | 705 hCoV-19/USA/NM-DOH-2020316957/2020\|EPI_ISL_732724\| | ···················· | ···················· | ····················· | ···················· | ···················· | ···················· |
| Y | 706 hCoV-19/USA/NM-DOH-2020317186/2020\|EPI_ISL_732725\| | ···················· | ···················· | ····················· | ···················· | ···················· | ···················· |
| Y | 707 hCoV-19/USA/NM-DOH-2020317191/2020\|EPI_ISL_732726\| | ···················· | ·A·················· | ····················· | ···················· | ···················· | ···················· |
| Y | 708 hCoV-19/USA/NM-DOH-2020317193/2020\|EPI_ISL_732727\| | ···················· | ···················· | ····················· | ···················· | ···················· | ···················· |
| Y | 709 hCoV-19/USA/NM-DOH-2020317452/2020\|EPI_ISL_732728\| | ···················· | ···················· | ····················· | ···················· | ···················· | ···················· |
| Y | 710 hCoV-19/USA/NM-DOH-2020317457/2020\|EPI_ISL_732729\| | ···················· | ·A·················· | ····················· | ···················· | ···················· | ···················· |
| Y | 711 hCoV-19/USA/NM-DOH-2020317582/2020\|EPI_ISL_732730\| | ···················· | ···················· | ····················· | ···················· | ···················· | ···················· |
| Y | 712 hCoV-19/USA/NM-DOH-2020317782/2020\|EPI_ISL_732731\| | ···················· | ···A················ | ····················· | ···················· | ···················· | ···················· |
| Y | 713 hCoV-19/USA/NM-DOH-2020318368/2020\|EPI_ISL_732732\| | ···················· | ···················· | ····················· | ···················· | ···················· | ···················· |
| Y | 714 hCoV-19/USA/NM-DOH-2020318478/2020\|EPI_ISL_732733\| | ···················· | ···················· | ····················· | ···················· | ···················· | ···················· |
| Y | 715 hCoV-19/USA/NM-DOH-2020318505/2020\|EPI_ISL_732734\| | ···················· | ·A·················· | ····················· | ···················· | ···················· | ···················· |
| Y | 716 hCoV-19/USA/NM-DOH-2020318519/2020\|EPI_ISL_732735\| | ···················· | ···················· | ····················· | ···················· | ···················· | ···················· |
| Y | 717 hCoV-19/USA/NM-DOH-2020318654/2020\|EPI_ISL_732736\| | ···················· | ···················· | ····················· | ···················· | ···················· | ···················· |
| Y | 718 hCoV-19/USA/NM-DOH-2020318667/2020\|EPI_ISL_732737\| | ···················· | ·A·················· | ····················· | ···················· | ···················· | ···················· |
| Y | 719 hCoV-19/USA/NM-DOH-2020318668/2020\|EPI_ISL_732738\| | ···················· | ···················· | ····················· | ···················· | ···················· | ···················· |
| Y | 720 hCoV-19/USA/NM-DOH-2020318741/2020\|EPI_ISL_732739\| | ···················· | ···················· | ····················· | ···················· | ···················· | ···················· |
| Y | 721 hCoV-19/USA/NM-DOH-2020318805/2020\|EPI_ISL_732740\| | ···················· | ···················· | ····················· | ···················· | ···················· | ···················· |
| Y | 722 hCoV-19/USA/NM-DOH-2020318937/2020\|EPI_ISL_732742\| | ···················· | ·A·················· | ····················· | ···················· | ···················· | ···················· |
| Y | 723 hCoV-19/USA/NM-DOH-2020318939/2020\|EPI_ISL_732743\| | ···················· | ···················· | ····················· | ···················· | ···················· | ···················· |
| Y | 724 hCoV-19/USA/NM-DOH-2020318940/2020\|EPI_ISL_732744\| | ···················· | ···················· | ····················· | ···················· | ···················· | ···················· |
| Y | 725 hCoV-19/USA/NM-DOH-2020319224/2020\|EPI_ISL_732746\| | ···················· | ···················· | ····················· | ···················· | ···················· | ···················· |
| Y | 726 hCoV-19/USA/NM-DOH-2020319240/2020\|EPI_ISL_732747\| | ···················· | ···················· | ····················· | ···················· | ···················· | ···················· |
| Y | 738 hCoV-19/Spain/RI-IBV-99010923/2020\|EPI_ISL_732762\| | ···················· | ···················· | ····················· | ···················· | ···················· | ···················· |
| Y | 739 hCoV-19/Spain/RI-IBV-99010924/2020\|EPI_ISL_732763\| | ···················· | ···················· | ····················· | ···················· | ···················· | ···················· |
| Y | 744 hCoV-19/Spain/RI-IBV-99010930/2020\|EPI_ISL_732768\| | ···················· | ···················· | ····················· | ···················· | ···················· | ···················· |
| Y | 754 hCoV-19/Spain/RI-IBV-99010953/2020\|EPI_ISL_732783\| | ···················· | ···················· | ····················· | ···················· | ··N················· | ···················· |
| Y | 758 hCoV-19/Spain/RI-IBV-99010960/2020\|EPI_ISL_732788\| | ···················· | ···················· | ····················· | ···················· | ···················· | ···················· |
| Y | 767 hCoV-19/England/QEUH-C57B94/2020\|EPI_ISL_736290\|20 | ···················· | ···················· | ····················· | ···················· | ···················· | ···················· |
| Y | 768 hCoV-19/England/MILK-C52C69/2020\|EPI_ISL_736299\|20 | ···················· | ···················· | ····················· | ···················· | ···················· | ···················· |
| Y | 769 hCoV-19/England/NOTT-11FA96/2020\|EPI_ISL_725604\|20 | ···················· | ···················· | ····················· | ···················· | ···················· | ···················· |
| Y | 770 hCoV-19/England/NOTT-120379/2020\|EPI_ISL_725607\|20 | ···················· | ···················· | ····················· | ···················· | ···················· | ···················· |
| Y | 771 hCoV-19/England/NOTT-120397/2020\|EPI_ISL_725608\|20 | ···················· | ···················· | ····················· | ···················· | ···················· | ···················· |
| Y | 772 hCoV-19/England/NOTT-12040D/2020\|EPI_ISL_725613\|20 | ···················· | ···················· | ····················· | ···················· | ···················· | ···················· |
| Y | 773 hCoV-19/England/NOTT-120555/2020\|EPI_ISL_725626\|20 | ···················· | ···················· | ····················· | ···················· | ···················· | ···················· |
| Y | 774 hCoV-19/England/NOTT-1205A0/2020\|EPI_ISL_725629\|20 | ···················· | ·A·················· | ····················· | ···················· | ···················· | ···················· |
| Y | 775 hCoV-19/England/NOTT-1205DD/2020\|EPI_ISL_725631\|20 | ···················· | ···················· | ····················· | ···················· | ···················· | ···················· |
| Y | 776 hCoV-19/England/NOTT-120616/2020\|EPI_ISL_725632\|20 | ···················· | ···················· | ····················· | ···················· | ···················· | ···················· |
| Y | 777 hCoV-19/England/NOTT-120643/2020\|EPI_ISL_725633\|20 | ···················· | ···················· | ····················· | ···················· | ···················· | ···················· |
| Y | 778 hCoV-19/England/NOTT-120661/2020\|EPI_ISL_725635\|20 | ···················· | ···················· | ····················· | ···················· | ···················· | ···················· |
| Y | 779 hCoV-19/England/NOTT-120713/2020\|EPI_ISL_725642\|20 | ···················· | ···················· | ····················· | ···················· | ···················· | ···················· |
| Y | 780 hCoV-19/England/NOTT-12076E/2020\|EPI_ISL_725643\|20 | ···················· | ···················· | ····················· | ···················· | ···················· | ···················· |
| Y | 781 hCoV-19/England/NOTT-1207AA/2020\|EPI_ISL_725646\|20 | ···················· | ···················· | ····················· | ···················· | ···················· | ···················· |
| Y | 782 hCoV-19/England/NOTT-120B44/2020\|EPI_ISL_725656\|20 | ···················· | ···················· | ····················· | ···················· | ···················· | ···················· |
| Y | 783 hCoV-19/England/NOTT-120C7E/2020\|EPI_ISL_725660\|20 | ···················· | ···················· | ····················· | ···················· | ···················· | ···················· |
| Y | 784 hCoV-19/Wales/PHWC-48C8F1/2020\|EPI_ISL_725670\|2020 | ···················· | ···················· | ····················· | ···················· | ···················· | ···················· |
| Y | 786 hCoV-19/Wales/PHWC-4925E2/2020\|EPI_ISL_725672\|2020 | ···················· | ···················· | ····················· | ···················· | ···················· | ···················· |
| Y | 787 hCoV-19/Wales/PHWC-4925F1/2020\|EPI_ISL_725673\|2020 | ···················· | ···················· | ····················· | ···················· | ···················· | ···················· |
| Y | 789 hCoV-19/Wales/PHWC-49261C/2020\|EPI_ISL_725675\|2020 | ···················· | ···················· | ····················· | ···················· | ···················· | ···················· |
| Y | 791 hCoV-19/Wales/PHWC-49263A/2020\|EPI_ISL_725677\|2020 | ···················· | ···················· | ····················· | ···················· | ···················· | ···················· |
| Y | 793 hCoV-19/Wales/PHWC-4978E4/2020\|EPI_ISL_726900\|2020 | ···················· | ···················· | ····················· | ···················· | ···················· | ···················· |
| Y | 794 hCoV-19/Wales/PHWC-4978F3/2020\|EPI_ISL_726901\|2020 | ···················· | ···················· | ····················· | ···················· | ···················· | ···················· |
| Y | 795 hCoV-19/Wales/PHWC-49790F/2020\|EPI_ISL_726902\|2020 | ···················· | ···················· | ····················· | ···················· | ···················· | ···················· |
| Y | 796 hCoV-19/Wales/PHWC-49791E/2020\|EPI_ISL_726903\|2020 | ···················· | ···················· | ····················· | ···················· | ···················· | ···················· |
| Y | 797 hCoV-19/Wales/PHWC-4979D2/2020\|EPI_ISL_726915\|2020 | ···················· | ···················· | ····················· | ···················· | ···················· | ···················· |
| Y | 798 hCoV-19/Wales/PHWC-4979E1/2020\|EPI_ISL_726916\|2020 | ···················· | ···················· | ····················· | ···················· | ···················· | ···················· |

| | | | | | | | |
|---|---|---|---|---|---|---|---|
| Y | 799 hCoV-19/Wales/PHWC-4979F0/2020\|EPI_ISL_726917\|2020 | ···················· | ···················· | ···················· | ···················· | ···················· | ···················· |
| Y | 800 hCoV-19/Wales/PHWC-497A0C/2020\|EPI_ISL_726918\|2020 | ···················· | ···················· | ···················· | ···················· | ···················· | ···················· |
| Y | 801 hCoV-19/Wales/PHWC-497A1B/2020\|EPI_ISL_726919\|2020 | ···················· | ···················· | ···················· | ···················· | ···················· | ···················· |
| Y | 803 hCoV-19/Wales/PHWC-497A93/2020\|EPI_ISL_726927\|2020 | ···················· | ···················· | ···················· | ···················· | ···················· | ···················· |
| Y | 804 hCoV-19/Wales/PHWC-497AA2/2020\|EPI_ISL_726928\|2020 | ···················· | ···················· | ···················· | ···················· | ···················· | ···················· |
| Y | 805 hCoV-19/Wales/PHWC-497AB1/2020\|EPI_ISL_726929\|2020 | ···················· | ···················· | ···················· | ···················· | ···················· | ···················· |
| Y | 806 hCoV-19/Wales/PHWC-497AC0/2020\|EPI_ISL_726930\|2020 | ···················· | ···················· | ···················· | ···················· | ···················· | ···················· |
| Y | 807 hCoV-19/Wales/PHWC-497ADF/2020\|EPI_ISL_726931\|2020 | ···················· | ···················· | ···················· | ···················· | ···················· | ···················· |
| Y | 808 hCoV-19/Wales/PHWC-497AEE/2020\|EPI_ISL_726932\|2020 | ···················· | ···················· | ···················· | ···················· | ···················· | ···················· |
| Y | 809 hCoV-19/Wales/PHWC-497AFD/2020\|EPI_ISL_726933\|2020 | ···················· | ···················· | ···················· | ···················· | ···················· | ···················· |
| Y | 811 hCoV-19/Wales/PHWC-497B18/2020\|EPI_ISL_726935\|2020 | ···················· | ···················· | ···················· | ···················· | ···················· | ···················· |
| Y | 812 hCoV-19/Wales/PHWC-497B27/2020\|EPI_ISL_726936\|2020 | ···················· | ···················· | ···················· | ···················· | ···················· | ···················· |
| Y | 813 hCoV-19/Wales/PHWC-497B54/2020\|EPI_ISL_726938\|2020 | ···················· | ···················· | ···················· | ···················· | ···················· | ···················· |
| Y | 814 hCoV-19/Wales/PHWC-497B72/2020\|EPI_ISL_726940\|2020 | ···················· | ···················· | ···················· | ···················· | ···················· | ···················· |
| Y | 815 hCoV-19/Wales/PHWC-497B90/2020\|EPI_ISL_726941\|2020 | ···················· | ···················· | ···················· | ···················· | ···················· | ···················· |
| Y | 816 hCoV-19/Wales/PHWC-497BAF/2020\|EPI_ISL_726942\|2020 | ···················· | ···················· | ···················· | ···················· | ···················· | ···················· |
| Y | 817 hCoV-19/Wales/PHWC-497BBE/2020\|EPI_ISL_726943\|2020 | ···················· | ···················· | ···················· | ···················· | ···················· | ···················· |
| Y | 818 hCoV-19/Wales/PHWC-497BCD/2020\|EPI_ISL_726944\|2020 | ···················· | ···················· | ···················· | ···················· | ···················· | ···················· |
| Y | 819 hCoV-19/Wales/PHWC-497BDC/2020\|EPI_ISL_726945\|2020 | ···················· | ···················· | ···················· | ···················· | ···················· | ···················· |
| Y | 820 hCoV-19/Wales/PHWC-497BEB/2020\|EPI_ISL_726946\|2020 | ···················· | ···················· | ···················· | ···················· | ···················· | ···················· |
| Y | 821 hCoV-19/Wales/PHWC-497BFA/2020\|EPI_ISL_726947\|2020 | ···················· | ···················· | ···················· | ···················· | ···················· | ···················· |
| Y | 822 hCoV-19/Wales/PHWC-497C06/2020\|EPI_ISL_726948\|2020 | ···················· | ···················· | ···················· | ···················· | ···················· | ···················· |
| Y | 825 hCoV-19/Wales/PHWC-497C51/2020\|EPI_ISL_726953\|2020 | ···················· | ···················· | ···················· | ···················· | ···················· | ···················· |
| Y | 826 hCoV-19/Wales/PHWC-497C60/2020\|EPI_ISL_726954\|2020 | ···················· | ·····A·············· | ···················· | ···················· | ···················· | ···················· |
| Y | 827 hCoV-19/Wales/PHWC-497C8E/2020\|EPI_ISL_726956\|2020 | ···················· | ···················· | ···················· | ···················· | ···················· | ···················· |
| Y | 828 hCoV-19/Wales/PHWC-497C9D/2020\|EPI_ISL_726957\|2020 | ···················· | ·····A·············· | ···················· | ···················· | ···················· | ···················· |
| Y | 830 hCoV-19/Wales/PHWC-497CBB/2020\|EPI_ISL_726959\|2020 | ···················· | ···················· | ···················· | ···················· | ···················· | ···················· |
| Y | 831 hCoV-19/Wales/PHWC-497CCA/2020\|EPI_ISL_726960\|2020 | ···················· | ·····A·············· | ···················· | ···················· | ···················· | ···················· |
| Y | 832 hCoV-19/Wales/PHWC-497CD9/2020\|EPI_ISL_726961\|2020 | ···················· | ···················· | ···················· | ···················· | ···················· | ···················· |
| Y | 833 hCoV-19/Wales/PHWC-497CF7/2020\|EPI_ISL_726963\|2020 | ···················· | ···················· | ···················· | ···················· | ···················· | ···················· |
| Y | 834 hCoV-19/Wales/PHWC-497D03/2020\|EPI_ISL_726964\|2020 | ···················· | ···················· | ···················· | ···················· | ···················· | ···················· |
| Y | 835 hCoV-19/Wales/PHWC-497D30/2020\|EPI_ISL_726967\|2020 | ···················· | ···················· | ···················· | ···················· | ···················· | ···················· |
| Y | 836 hCoV-19/Wales/PHWC-497D4F/2020\|EPI_ISL_726968\|2020 | ···················· | ···················· | ···················· | ···················· | ···················· | ···················· |
| Y | 837 hCoV-19/Wales/PHWC-497D5E/2020\|EPI_ISL_726969\|2020 | ···················· | ···················· | ···················· | ···················· | ···················· | ···················· |
| Y | 838 hCoV-19/Wales/PHWC-497D6D/2020\|EPI_ISL_726970\|2020 | ···················· | ···················· | ···················· | ···················· | ···················· | ···················· |
| Y | 839 hCoV-19/Wales/PHWC-497D8B/2020\|EPI_ISL_726972\|2020 | ···················· | ···················· | ···················· | ···················· | ···················· | ···················· |
| Y | 840 hCoV-19/Wales/PHWC-497D9A/2020\|EPI_ISL_726973\|2020 | ···················· | ···················· | ···················· | ···················· | ···················· | ···················· |
| Y | 841 hCoV-19/Wales/PHWC-497DA9/2020\|EPI_ISL_726974\|2020 | ···················· | ···················· | ···················· | ···················· | ···················· | ···················· |
| Y | 842 hCoV-19/Wales/PHWC-497DB8/2020\|EPI_ISL_726975\|2020 | ···················· | ···················· | ···················· | ···················· | ···················· | ···················· |
| Y | 843 hCoV-19/Wales/PHWC-497DC7/2020\|EPI_ISL_726976\|2020 | ···················· | ···················· | ···················· | ···················· | ···················· | ···················· |
| Y | 844 hCoV-19/Wales/PHWC-497DD6/2020\|EPI_ISL_726977\|2020 | ···················· | ···················· | ···················· | ···················· | ···················· | ···················· |
| Y | 845 hCoV-19/Wales/PHWC-497DE5/2020\|EPI_ISL_726978\|2020 | ···················· | ···················· | ···················· | ···················· | ···················· | ···················· |
| Y | 846 hCoV-19/Wales/PHWC-497DF4/2020\|EPI_ISL_726979\|2020 | ···················· | ···················· | ···················· | ···················· | ···················· | ···················· |
| Y | 847 hCoV-19/Wales/PHWC-497E00/2020\|EPI_ISL_726980\|2020 | ···················· | ···················· | ···················· | ···················· | ···················· | ···················· |
| Y | 848 hCoV-19/Wales/PHWC-497E1F/2020\|EPI_ISL_726981\|2020 | ···················· | ···················· | ···················· | ···················· | ···················· | ···················· |
| Y | 849 hCoV-19/Wales/PHWC-497E2E/2020\|EPI_ISL_726982\|2020 | ···················· | ···················· | ···················· | ···················· | ···················· | ···················· |
| Y | 850 hCoV-19/Wales/PHWC-497E3D/2020\|EPI_ISL_726983\|2020 | ···················· | ···················· | ···················· | ···················· | ···················· | ···················· |
| Y | 851 hCoV-19/Wales/PHWC-497E4C/2020\|EPI_ISL_726984\|2020 | ···················· | ···················· | ···················· | ···················· | ···················· | ···················· |
| Y | 852 hCoV-19/Wales/PHWC-497E5B/2020\|EPI_ISL_726985\|2020 | ···················· | ···················· | ···················· | ···················· | ···················· | ···················· |
| Y | 853 hCoV-19/Wales/PHWC-497E6A/2020\|EPI_ISL_726986\|2020 | ···················· | ···················· | ···················· | ···················· | ···················· | ···················· |
| Y | 854 hCoV-19/Wales/PHWC-497E79/2020\|EPI_ISL_726987\|2020 | ···················· | ···················· | ···················· | ···················· | ···················· | ···················· |
| Y | 855 hCoV-19/Wales/PHWC-497E88/2020\|EPI_ISL_726988\|2020 | ···················· | ···················· | ···················· | ···················· | ···················· | ···················· |
| Y | 856 hCoV-19/Wales/PHWC-497E97/2020\|EPI_ISL_726989\|2020 | ···················· | ···················· | ···················· | ···················· | ···················· | ···················· |
| Y | 857 hCoV-19/Wales/PHWC-497EA6/2020\|EPI_ISL_726990\|2020 | ···················· | ···················· | ···················· | ···················· | ···················· | ···················· |
| Y | 858 hCoV-19/Wales/PHWC-497EB5/2020\|EPI_ISL_726991\|2020 | ···················· | ···················· | ···················· | ···················· | ···················· | ···················· |
| Y | 859 hCoV-19/Wales/PHWC-497EC4/2020\|EPI_ISL_726992\|2020 | ···················· | ···················· | ···················· | ···················· | ···················· | ···················· |
| Y | 860 hCoV-19/Wales/PHWC-497ED3/2020\|EPI_ISL_726993\|2020 | ···················· | ···················· | ···················· | ···················· | ···················· | ···················· |
| Y | 861 hCoV-19/Wales/PHWC-497EE2/2020\|EPI_ISL_726994\|2020 | ···················· | ···················· | ···················· | ···················· | ···················· | ···················· |
| Y | 862 hCoV-19/Wales/PHWC-497EF1/2020\|EPI_ISL_726995\|2020 | ···················· | ···················· | ···················· | ···················· | ···················· | ···················· |
| Y | 863 hCoV-19/Wales/PHWC-497F0D/2020\|EPI_ISL_726996\|2020 | ···················· | ···················· | ···················· | ···················· | ···················· | ···················· |
| Y | 864 hCoV-19/Wales/PHWC-497F1C/2020\|EPI_ISL_726997\|2020 | ···················· | ···················· | ···················· | ···················· | ···················· | ···················· |
| Y | 865 hCoV-19/Wales/PHWC-497F3A/2020\|EPI_ISL_726999\|2020 | ···················· | ···················· | ···················· | ···················· | ···················· | ···················· |
| Y | 866 hCoV-19/Switzerland/SO-ETHZ-400302/2020\|EPI_ISL_72 | ···················· | ···················· | ···················· | ···················· | ···················· | ···················· |
| Y | 867 hCoV-19/Switzerland/SO-ETHZ-400305/2020\|EPI_ISL_72 | ···················· | ···················· | ···················· | ···················· | ···················· | ···················· |
| Y | 868 hCoV-19/Switzerland/SO-ETHZ-400307/2020\|EPI_ISL_72 | ···················· | ···················· | ···················· | ···················· | ···················· | ···················· |
| Y | 869 hCoV-19/Switzerland/SO-ETHZ-400308/2020\|EPI_ISL_72 | ···················· | ···················· | ···················· | ···················· | ···················· | ···················· |
| Y | 870 hCoV-19/Switzerland/SO-ETHZ-400309/2020\|EPI_ISL_72 | ···················· | ···················· | ···················· | ···················· | ···················· | ···················· |
| Y | 871 hCoV-19/Switzerland/ZH-ETHZ-400331/2020\|EPI_ISL_72 | ···················· | ···················· | ···················· | ···················· | ···················· | ···················· |
| Y | 872 hCoV-19/Switzerland/SH-ETHZ-400332/2020\|EPI_ISL_72 | ···················· | ···················· | ···················· | ···················· | ···················· | ···················· |
| Y | 873 hCoV-19/Switzerland/ZH-ETHZ-400333/2020\|EPI_ISL_72 | ···················· | ···················· | ···················· | ···················· | ···················· | ···················· |
| Y | 874 hCoV-19/Switzerland/ZH-ETHZ-400335/2020\|EPI_ISL_72 | ···················· | ···················· | ···················· | ···················· | ···················· | ···················· |
| Y | 875 hCoV-19/Switzerland/ZH-ETHZ-400336/2020\|EPI_ISL_72 | ···················· | ···················· | ···················· | ···················· | ···················· | ···················· |
| Y | 876 hCoV-19/Switzerland/ZH-ETHZ-400337/2020\|EPI_ISL_72 | ···················· | ···················· | ···················· | ···················· | ···················· | ···················· |
| Y | 877 hCoV-19/Switzerland/ZH-ETHZ-400338/2020\|EPI_ISL_72 | ···················· | ···················· | ···················· | ···················· | ···················· | ···················· |
| Y | 878 hCoV-19/Switzerland/BE-ETHZ-400340/2020\|EPI_ISL_72 | ···················· | ···················· | ···················· | ···················· | ···················· | ···················· |
| Y | 879 hCoV-19/Switzerland/BE-ETHZ-400342/2020\|EPI_ISL_72 | ···················· | ···················· | ···················· | ···················· | ···················· | ···················· |
| Y | 880 hCoV-19/Switzerland/BE-ETHZ-400344/2020\|EPI_ISL_72 | ···················· | ···················· | ···················· | ···················· | ···················· | ···················· |
| Y | 881 hCoV-19/Switzerland/ZH-ETHZ-400345/2020\|EPI_ISL_72 | ···················· | ···················· | ···················· | ···················· | ·····K·············· | ···················· |
| Y | 882 hCoV-19/Switzerland/ZH-ETHZ-400346/2020\|EPI_ISL_72 | ···················· | ···················· | ···················· | ···················· | ···················· | ···················· |
| Y | 883 hCoV-19/Switzerland/BE-ETHZ-400347/2020\|EPI_ISL_72 | ···················· | ···················· | ···················· | ···················· | ···················· | ···················· |

Y 884 hCoV-19/Switzerland/BE-ETHZ-400348/2020|EPI_ISL_72 ···················· ···················· ···················· ···················· ···················· ····················
Y 885 hCoV-19/Switzerland/BE-ETHZ-400349/2020|EPI_ISL_72 ···················· ···················· ···················· ···················· ···················· ····················
Y 886 hCoV-19/Switzerland/BL-ETHZ-400390/2020|EPI_ISL_72 ···················· ···················· ···················· ···················· ···················· ····················
Y 887 hCoV-19/Switzerland/BL-ETHZ-400391/2020|EPI_ISL_72 ···················· ···················· ···················· ···················· ···················· ····················
Y 888 hCoV-19/Switzerland/AG-ETHZ-400396/2020|EPI_ISL_72 ···················· ···················· ···················· ···················· ···················· ····················
Y 889 hCoV-19/USA/CA-ALSR-4800/2020|EPI_ISL_730150|2020- ···················· ·A·················· ···················· ···················· ···················· ····················
Y 890 hCoV-19/USA/CA-ALSR-4801/2020|EPI_ISL_730151|2020- ···················· ·A·················· ···················· ···················· ···················· ····················
Y 891 hCoV-19/USA/CA-ALSR-4802/2020|EPI_ISL_730152|2020- ···················· ·A·················· ···················· ···················· ···················· ····················
Y 892 hCoV-19/USA/CA-ALSR-4811/2020|EPI_ISL_730155|2020- ···················· ···················· ···················· ···················· ···················· ····················
Y 893 hCoV-19/USA/CA-ALSR-4814/2020|EPI_ISL_730156|2020- ···················· ···················· ···················· ···················· ···················· ····················
Y 894 hCoV-19/USA/CA-ALSR-4816/2020|EPI_ISL_730158|2020- ···················· ···················· ···················· ···················· ···················· ····················
Y 895 hCoV-19/USA/CA-ALSR-4818/2020|EPI_ISL_730159|2020- ···················· ·A·················· ···················· ···················· ···················· ····················
Y 896 hCoV-19/USA/CA-ALSR-4820/2020|EPI_ISL_730160|2020- ···················· ···················· ···················· ···················· ···················· ····················
Y 897 hCoV-19/USA/CA-ALSR-4821/2020|EPI_ISL_730161|2020- ···················· ···················· ···················· ···················· ···················· ····················
Y 898 hCoV-19/USA/CA-ALSR-4822/2020|EPI_ISL_730162|2020- ···················· ···················· ···················· ···················· ···················· ····················
Y 899 hCoV-19/USA/CA-ALSR-4825/2020|EPI_ISL_730163|2020- ···················· ···················· ···················· ···················· ···················· ····················
Y 900 hCoV-19/USA/CA-ALSR-4826/2020|EPI_ISL_730164|2020- ···················· ···················· ···················· ···················· ···················· ····················
Y 901 hCoV-19/USA/CA-ALSR-4828/2020|EPI_ISL_730165|2020- ···················· ···················· ···················· ···················· ···················· ····················
Y 902 hCoV-19/USA/CA-ALSR-4830/2020|EPI_ISL_730166|2020- ···················· ···················· ···················· ···················· ···················· ····················
Y 903 hCoV-19/USA/CA-ALSR-4831/2020|EPI_ISL_730167|2020- ···················· ···················· ···················· ···················· ···················· ····················
Y 904 hCoV-19/USA/CA-ALSR-4834/2020|EPI_ISL_730168|2020- ···················· ···················· ···················· ···················· ···················· ····················
Y 905 hCoV-19/USA/CA-ALSR-4835/2020|EPI_ISL_730169|2020- ···················· ···················· ···················· ···················· ···················· ····················
Y 906 hCoV-19/USA/CA-ALSR-4837/2020|EPI_ISL_730170|2020- ···················· ···················· ···················· ···················· ···················· ····················
Y 907 hCoV-19/USA/CA-ALSR-4839/2020|EPI_ISL_730171|2020- ···················· ···················· ···················· ···················· ···················· ····················
Y 908 hCoV-19/USA/CA-ALSR-4841/2020|EPI_ISL_730172|2020- ···················· ···················· ···················· ···················· ···················· ····················
Y 909 hCoV-19/USA/CA-ALSR-4842/2020|EPI_ISL_730173|2020- ···················· ···················· ···················· ···················· ···················· ····················
Y 910 hCoV-19/USA/CA-ALSR-4846/2020|EPI_ISL_730175|2020- ···················· ·A·················· ···················· ···················· ···················· ····················
Y 911 hCoV-19/USA/CA-ALSR-4848/2020|EPI_ISL_730176|2020- ···················· ···················· ···················· ···················· ···················· ····················
Y 912 hCoV-19/USA/CA-ALSR-4849/2020|EPI_ISL_730177|2020- ···················· ·A·················· ···················· ···················· ···················· ····················
Y 913 hCoV-19/USA/CA-ALSR-4854/2020|EPI_ISL_730178|2020- ···················· ···················· ···················· ···················· ···················· ····················
Y 914 hCoV-19/USA/CA-ALSR-4856/2020|EPI_ISL_730179|2020- ···················· ···················· ···················· ···················· ···················· ····················
Y 915 hCoV-19/USA/CA-ALSR-4871/2020|EPI_ISL_730190|2020- ···················· ···················· ···················· ···················· ···················· ····················
Y 916 hCoV-19/USA/CA-ALSR-4872/2020|EPI_ISL_730191|2020- ···················· ·A·················· ···················· ···················· ···················· ····················
Y 917 hCoV-19/USA/CA-ALSR-4873/2020|EPI_ISL_730192|2020- ···················· ···················· ···················· ···················· ···················· ····················
Y 918 hCoV-19/USA/CA-ALSR-4874/2020|EPI_ISL_730193|2020- ···················· ·A·················· ···················· ···················· ···················· ····················
Y 919 hCoV-19/USA/CA-ALSR-4877/2020|EPI_ISL_730194|2020- ···················· ···················· ···················· ···················· ···················· ····················
Y 920 hCoV-19/USA/CA-ALSR-4879/2020|EPI_ISL_730195|2020- ···················· ·A·················· ···················· ···················· ···················· ····················
Y 921 hCoV-19/USA/CA-ALSR-4885/2020|EPI_ISL_730196|2020- ···················· ···················· ···················· ···················· ···················· ····················
Y 922 hCoV-19/Mexico/BCN-ALSR-4888/2020|EPI_ISL_730197|2 ···················· ···················· ···················· ···················· ···················· ····················
Y 923 hCoV-19/Mexico/BCN-ALSR-4894/2020|EPI_ISL_730199|2 ···················· ·A·················· ···················· ···················· ···················· ····················
Y 924 hCoV-19/England/ALDP-C3A5E4/2020|EPI_ISL_731481|20 ···················· ···················· ···················· ···················· ···················· ····················
Y 925 hCoV-19/England/ALDP-C39239/2020|EPI_ISL_731483|20 ···················· ···················· ···················· ···················· ···················· ····················
Y 926 hCoV-19/England/QEUH-C30BDD/2020|EPI_ISL_731484|20 ···················· ···················· ···················· ···················· ···················· ····················
Y 927 hCoV-19/England/QEUH-C30AC1/2020|EPI_ISL_731485|20 ···················· ···················· ···················· ···················· ···················· ····················
Y 928 hCoV-19/England/QEUH-C35F63/2020|EPI_ISL_731486|20 ···················· ···················· ···················· ···················· ···················· ····················
Y 929 hCoV-19/England/ALDP-C39275/2020|EPI_ISL_731487|20 ···················· ···················· ···················· ···················· ···················· ····················
Y 930 hCoV-19/England/QEUH-C31037/2020|EPI_ISL_731488|20 ···················· ···················· ···················· ···················· ···················· ····················
Y 931 hCoV-19/England/ALDP-C39327/2020|EPI_ISL_731489|20 ···················· ···················· ···················· ···················· ······T············· ····················
Y 932 hCoV-19/England/QEUH-C310CE/2020|EPI_ISL_731491|20 ···················· ···················· ···················· ···················· ···················· ····················
Y 933 hCoV-19/England/QEUH-C34C30/2020|EPI_ISL_731493|20 ···················· ···················· ···················· ···················· ···················· ····················
Y 934 hCoV-19/England/ALDP-C39345/2020|EPI_ISL_731495|20 ···················· ···················· ···················· ···················· ···················· ····················
Y 935 hCoV-19/England/QEUH-C311DA/2020|EPI_ISL_731496|20 ···················· ···················· ···················· ···················· ···················· ····················
Y 936 hCoV-19/England/ALDP-C39855/2020|EPI_ISL_731498|20 ···················· ···················· ···················· ···················· ······T············· ····················
Y 937 hCoV-19/England/QEUH-C31529/2020|EPI_ISL_731499|20 ···················· ···················· ···················· ···················· ···················· ····················
Y 938 hCoV-19/Belgium/rega-09221024/2020|EPI_ISL_735021| ···················· ···················· ···················· ···················· ···················· ····················
Y 939 hCoV-19/Belgium/rega-09221064/2020|EPI_ISL_735022| ···················· ···················· ···················· ···················· ···················· ····················
Y 940 hCoV-19/Belgium/rega-09221065/2020|EPI_ISL_735023| ···················· ···················· ···················· ···················· ···················· ····················
Y 942 hCoV-19/Belgium/rega-09221235/2020|EPI_ISL_735025| ···················· ···················· ···················· ···················· ···················· ····················
Y 943 hCoV-19/Belgium/rega-09231003/2020|EPI_ISL_735026| ···················· ···················· ···················· ···················· ···················· ····················
Y 944 hCoV-19/Belgium/rega-09231004/2020|EPI_ISL_735027| ···················· ···················· ···················· ···················· ···················· ····················
Y 945 hCoV-19/Belgium/rega-09231005/2020|EPI_ISL_735028| ···················· ···················· ···················· ···················· ···················· ····················
Y 946 hCoV-19/Belgium/rega-09231006/2020|EPI_ISL_735029| ···················· ···················· ···················· ···················· ···················· ····················
Y 947 hCoV-19/Belgium/rega-09231007/2020|EPI_ISL_735030| ···················· ···················· ···················· ···················· ···················· ····················
Y 948 hCoV-19/Belgium/rega-09231008/2020|EPI_ISL_735031| ···················· ···················· ···················· ···················· ···················· ····················
Y 949 hCoV-19/Belgium/rega-09231023/2020|EPI_ISL_735032| ···················· ···················· ···················· ···················· ···················· ····················
Y 950 hCoV-19/Belgium/rega-09231025/2020|EPI_ISL_735033| ···················· ···················· ···················· ···················· ···················· ····················
Y 951 hCoV-19/Belgium/rega-09231187/2020|EPI_ISL_735034| ···················· ···················· ···················· ···················· ···················· ····················
Y 952 hCoV-19/Belgium/rega-09231248/2020|EPI_ISL_735035| ···················· ·A·················· ···················· ···················· ···················· ····················
Y 953 hCoV-19/Belgium/rega-09241212/2020|EPI_ISL_735036| ···················· ···················· ···················· ···················· ···················· ····················
Y 954 hCoV-19/Belgium/rega-09241218/2020|EPI_ISL_735037| ···················· ···················· ···················· ···················· ···················· ····················
Y 955 hCoV-19/Belgium/rega-09241224/2020|EPI_ISL_735038| ···················· ···················· ···················· ···················· ···················· ····················
Y 956 hCoV-19/Belgium/rega-09241230/2020|EPI_ISL_735039| ···················· ···················· ···················· ···················· ···················· ····················
Y 957 hCoV-19/England/QEUH-C5456C/2020|EPI_ISL_736304|20 ···················· ···················· ···················· ···················· ···················· ····················
Y 958 hCoV-19/England/MILK-C5263E/2020|EPI_ISL_736308|20 ···················· ···················· ···················· ···················· ···················· ····················
Y 959 hCoV-19/England/QEUH-C53F14/2020|EPI_ISL_736309|20 ···················· ···················· ···················· ···················· ···················· ···A················
Y 960 hCoV-19/England/QEUH-C53F7E/2020|EPI_ISL_736312|20 ···················· ···················· ···················· ···················· ···················· ····················
Y 961 hCoV-19/England/QEUH-C57BA3/2020|EPI_ISL_736313|20 ···················· ···················· ···················· ···················· ···················· ····················
Y 962 hCoV-19/England/MILK-C52E54/2020|EPI_ISL_736314|20 ···················· ···················· ···················· ···················· ···················· ····················
Y 963 hCoV-19/England/QEUH-C57C0A/2020|EPI_ISL_736315|20 ···················· ···················· ···················· ···················· ···················· ····················
Y 964 hCoV-19/England/QEUH-C57A3D/2020|EPI_ISL_736316|20 ···················· ···················· ···················· ···················· ···················· ····················

| | | | | | | | | |
|---|---|---|---|---|---|---|---|---|
| Y | 965 | hCoV-19/England/MILK-C52ECD/2020\|EPI_ISL_736317\|20 | ···················· | ···················· | ···················· | ···················· | ···················· | ···················· |
| Y | 966 | hCoV-19/England/QEUH-C53ECC/2020\|EPI_ISL_736321\|20 | ···················· | ·A·················· | ···················· | ···················· | ···················· | ···················· |
| Y | 967 | hCoV-19/England/QEUH-C542B0/2020\|EPI_ISL_736325\|20 | ···················· | ···················· | ···················· | ···················· | ···················· | ···················· |
| Y | 968 | hCoV-19/England/MILK-C52C78/2020\|EPI_ISL_736329\|20 | ···················· | ···················· | ···················· | ···················· | ···················· | ···················· |
| Y | 969 | hCoV-19/England/QEUH-C54177/2020\|EPI_ISL_736352\|20 | ···················· | ···················· | ···················· | ···················· | ······T············· | ···················· |
| Y | 970 | hCoV-19/England/QEUH-C5453F/2020\|EPI_ISL_736355\|20 | ···················· | ···················· | ···················· | ···················· | ···················· | ···················· |
| Y | 971 | hCoV-19/England/QEUH-C57AA6/2020\|EPI_ISL_736367\|20 | ···················· | ···················· | ···················· | ···················· | ···················· | ···················· |
| Y | 972 | hCoV-19/Switzerland/BE-ETHZ-410798/2020\|EPI_ISL_73 | ···················· | ···················· | ···················· | ···················· | ···················· | ···················· |
| Y | 975 | hCoV-19/Switzerland/VS-ETHZ-410467/2020\|EPI_ISL_73 | ···················· | ···················· | ···················· | ···················· | ···················· | ···················· |
| Y | 976 | hCoV-19/Switzerland/SO-ETHZ-410336/2020\|EPI_ISL_73 | ···················· | ···················· | ···················· | ···················· | ···················· | ···················· |
| Y | 977 | hCoV-19/Switzerland/VS-ETHZ-410006/2020\|EPI_ISL_73 | ···················· | ···················· | ···················· | ···················· | ···················· | ···················· |
| Y | 978 | hCoV-19/Switzerland/BL-ETHZ-410810/2020\|EPI_ISL_73 | ···················· | ···················· | ···················· | ···················· | ···················· | ···················· |
| Y | 979 | hCoV-19/Switzerland/BE-ETHZ-320204/2020\|EPI_ISL_73 | ···················· | ···················· | ···················· | ···················· | ···················· | ···················· |
| Y | 980 | hCoV-19/Switzerland/SO-ETHZ-410954/2020\|EPI_ISL_73 | ···················· | ···················· | ···················· | ···················· | ···················· | ···················· |
| Y | 981 | hCoV-19/Switzerland/VS-ETHZ-410644/2020\|EPI_ISL_73 | ···················· | ···················· | ···················· | ···················· | ···················· | ···················· |
| Y | 982 | hCoV-19/Switzerland/ZH-ETHZ-410081/2020\|EPI_ISL_73 | ···················· | ···················· | ···················· | ···················· | ···················· | ···················· |
| Y | 983 | hCoV-19/Switzerland/ZH-ETHZ-410089/2020\|EPI_ISL_73 | ···················· | ···················· | ···················· | ···················· | ···················· | ···················· |
| Y | 984 | hCoV-19/Switzerland/ZH-ETHZ-410090/2020\|EPI_ISL_73 | ···················· | ···················· | ···················· | ···················· | ···················· | ···················· |
| Y | 985 | hCoV-19/Switzerland/ZH-ETHZ-410091/2020\|EPI_ISL_73 | ···················· | ···················· | ···················· | ···················· | ···················· | ···················· |
| Y | 986 | hCoV-19/Switzerland/BE-ETHZ-410565/2020\|EPI_ISL_73 | ···················· | ···················· | ···················· | ···················· | ···················· | ···················· |
| Y | 987 | hCoV-19/Switzerland/SH-ETHZ-410915/2020\|EPI_ISL_73 | ···················· | ···················· | ···················· | ···················· | ···················· | ···················· |
| Y | 988 | hCoV-19/Switzerland/SH-ETHZ-410916/2020\|EPI_ISL_73 | ···················· | ···················· | ···················· | ···················· | ···················· | ···················· |
| Y | 989 | hCoV-19/Switzerland/VS-ETHZ-411035/2020\|EPI_ISL_73 | ···················· | ···················· | ···················· | ···················· | ···················· | ···················· |
| Y | 990 | hCoV-19/Switzerland/VS-ETHZ-410082/2020\|EPI_ISL_73 | ···················· | ···················· | ···················· | ···················· | ···················· | ···················· |
| Y | 991 | hCoV-19/Switzerland/VS-ETHZ-410426/2020\|EPI_ISL_73 | ···················· | ···················· | ···················· | ···················· | ···················· | ···················· |
| Y | 992 | hCoV-19/Switzerland/VS-ETHZ-410706/2020\|EPI_ISL_73 | ···················· | ···················· | ···················· | ···················· | ···················· | ···················· |
| Y | 993 | hCoV-19/Switzerland/un-ETHZ-1/2020\|EPI_ISL_737647\| | ···················· | ···················· | ···················· | ···················· | ···················· | ···················· |
| Y | 994 | hCoV-19/Switzerland/un-ETHZ-410562/2020\|EPI_ISL_73 | ···················· | ···················· | ···················· | ···················· | ···················· | ···················· |
| Y | 995 | hCoV-19/Switzerland/AG-ETHZ-410033/2020\|EPI_ISL_73 | ···················· | ·A·················· | ···················· | ···················· | ···················· | ··A················· |
| Y | 996 | hCoV-19/Switzerland/AG-ETHZ-410041/2020\|EPI_ISL_73 | ···················· | ···················· | ···················· | ···················· | ···················· | ···················· |
| Y | 997 | hCoV-19/Switzerland/AG-ETHZ-410042/2020\|EPI_ISL_73 | ···················· | ·A·················· | ···················· | ···················· | ···················· | ··A················· |
| Y | 998 | hCoV-19/Switzerland/AG-ETHZ-410077/2020\|EPI_ISL_73 | ···················· | ···················· | ···················· | ···················· | ···················· | ···················· |
| Y | 999 | hCoV-19/Switzerland/AG-ETHZ-410114/2020\|EPI_ISL_73 | ···················· | ···················· | ···················· | ···················· | ···················· | ···················· |
| Y | 1001 | hCoV-19/Switzerland/AG-ETHZ-410119/2020\|EPI_ISL_73 | ···················· | ···················· | ···················· | ···················· | ···················· | ···················· |
| Y | 1002 | hCoV-19/Switzerland/AG-ETHZ-410186/2020\|EPI_ISL_73 | ···················· | ···················· | ···················· | ···················· | ···················· | ···················· |
| Y | 1004 | hCoV-19/Switzerland/AG-ETHZ-410268/2020\|EPI_ISL_73 | ···················· | ···················· | ···················· | ···················· | ···················· | ···················· |
| Y | 1005 | hCoV-19/Switzerland/AG-ETHZ-410273/2020\|EPI_ISL_73 | ···················· | ···················· | ···················· | ···················· | ···················· | ···················· |
| Y | 1006 | hCoV-19/Switzerland/BL-ETHZ-410961/2020\|EPI_ISL_73 | ···················· | ···················· | ···················· | ···················· | ···················· | ···················· |
| Y | 1007 | hCoV-19/Switzerland/BE-ETHZ-320191/2020\|EPI_ISL_73 | ···················· | ···················· | ···················· | ···················· | ···················· | ···················· |
| Y | 1008 | hCoV-19/Switzerland/BE-ETHZ-320195/2020\|EPI_ISL_73 | ···················· | ···················· | ···················· | ···················· | ···················· | ···················· |
| Y | 1009 | hCoV-19/Switzerland/BE-ETHZ-320197/2020\|EPI_ISL_73 | ···················· | ···················· | ···················· | ···················· | ···················· | ···················· |
| Y | 1011 | hCoV-19/Switzerland/BE-ETHZ-320208/2020\|EPI_ISL_73 | ···················· | ···················· | ···················· | ···················· | ······R············· | ···················· |
| Y | 1013 | hCoV-19/Switzerland/BE-ETHZ-320581/2020\|EPI_ISL_73 | ···················· | ···················· | ···················· | ···················· | ···················· | ···················· |
| Y | 1014 | hCoV-19/Switzerland/BE-ETHZ-320582/2020\|EPI_ISL_73 | ···················· | ···················· | ···················· | ···················· | ···················· | ···················· |
| Y | 1015 | hCoV-19/Switzerland/BE-ETHZ-320588/2020\|EPI_ISL_73 | ···················· | ···················· | ···················· | ···················· | ···················· | ···················· |
| Y | 1017 | hCoV-19/Switzerland/BE-ETHZ-321079/2020\|EPI_ISL_73 | ···················· | ···················· | ···················· | ···················· | ···················· | ···················· |
| Y | 1018 | hCoV-19/Switzerland/BE-ETHZ-321080/2020\|EPI_ISL_73 | ···················· | ···················· | ···················· | ···················· | ···················· | ···················· |
| Y | 1019 | hCoV-19/Switzerland/BE-ETHZ-321081/2020\|EPI_ISL_73 | ···················· | ···················· | ···················· | ···················· | ···················· | ···················· |
| Y | 1020 | hCoV-19/Switzerland/BE-ETHZ-321082/2020\|EPI_ISL_73 | ···················· | ···················· | ···················· | ···················· | ···················· | ···················· |
| Y | 1021 | hCoV-19/Switzerland/BE-ETHZ-321083/2020\|EPI_ISL_73 | ···················· | ···················· | ···················· | ···················· | ···················· | ···················· |
| Y | 1022 | hCoV-19/Switzerland/BE-ETHZ-321084/2020\|EPI_ISL_73 | ···················· | ···················· | ···················· | ···················· | ···················· | ···················· |
| Y | 1023 | hCoV-19/Switzerland/BE-ETHZ-321085/2020\|EPI_ISL_73 | ···················· | ···················· | ···················· | ···················· | ···················· | ···················· |
| Y | 1024 | hCoV-19/Switzerland/BE-ETHZ-321086/2020\|EPI_ISL_73 | ···················· | ···················· | ···················· | ···················· | ···················· | ···················· |
| Y | 1025 | hCoV-19/Wales/PHWC-492685/2020\|EPI_ISL_725681\|2020 | ···················· | ···················· | ···················· | ···················· | ···················· | ···················· |
| Y | 1026 | hCoV-19/Wales/PHWC-492694/2020\|EPI_ISL_725682\|2020 | ···················· | ···················· | ···················· | ···················· | ···················· | ···················· |
| Y | 1028 | hCoV-19/Wales/PHWC-4926D0/2020\|EPI_ISL_725686\|2020 | ···················· | ···················· | ···················· | ···················· | ···················· | ···················· |
| Y | 1029 | hCoV-19/Wales/PHWC-4926EF/2020\|EPI_ISL_725687\|2020 | ···················· | ···················· | ···················· | ···················· | ···················· | ···················· |
| Y | 1031 | hCoV-19/Wales/PHWC-492728/2020\|EPI_ISL_725691\|2020 | ···················· | ···················· | ···················· | ···················· | ···················· | ···················· |
| Y | 1033 | hCoV-19/Wales/PHWC-492755/2020\|EPI_ISL_725694\|2020 | ···················· | ···················· | ···················· | ···················· | ···················· | ···················· |
| Y | 1034 | hCoV-19/Wales/PHWC-492773/2020\|EPI_ISL_725696\|2020 | ···················· | ···················· | ···················· | ···················· | ···················· | ···················· |
| Y | 1035 | hCoV-19/Wales/PHWC-492791/2020\|EPI_ISL_725698\|2020 | ···················· | ···················· | ···················· | ···················· | ···················· | ···················· |
| Y | 1036 | hCoV-19/Wales/PHWC-4927A0/2020\|EPI_ISL_725699\|2020 | ···················· | ···················· | ···················· | ···················· | ···················· | ···················· |
| Y | 1037 | hCoV-19/Mexico/BCN-ALSR-4928/2020\|EPI_ISL_730217\|2 | ···················· | ·A·················· | ···················· | ···················· | ···················· | ···················· |
| Y | 1038 | hCoV-19/Mexico/BCN-ALSR-4929/2020\|EPI_ISL_730218\|2 | ···················· | ·A·················· | ···················· | ···················· | ···················· | ···················· |
| Y | 1039 | hCoV-19/England/ALDP-C394CA/2020\|EPI_ISL_731500\|20 | ···················· | ···················· | ···················· | ···················· | ···················· | ···················· |
| Y | 1040 | hCoV-19/England/QEUH-C30A0D/2020\|EPI_ISL_731501\|20 | ···················· | ···················· | ···················· | ···················· | ···················· | ···················· |
| Y | 1041 | hCoV-19/England/ALDP-C3AB6A/2020\|EPI_ISL_731502\|20 | ···················· | ···················· | ···················· | ···················· | ···················· | ···················· |
| Y | 1042 | hCoV-19/England/ALDP-C392B1/2020\|EPI_ISL_731504\|20 | ···················· | ···················· | ···················· | ···················· | ···················· | ···················· |
| Y | 1043 | hCoV-19/England/ALDP-C3A1D1/2020\|EPI_ISL_731506\|20 | ···················· | ···················· | ···················· | ···················· | ···················· | ···················· |
| Y | 1044 | hCoV-19/England/QEUH-C30C25/2020\|EPI_ISL_731507\|20 | ···················· | ···················· | ···················· | ···················· | ···················· | ···················· |
| Y | 1045 | hCoV-19/England/QEUH-C30AB2/2020\|EPI_ISL_731508\|20 | ···················· | ···················· | ···················· | ···················· | ···················· | ···················· |
| Y | 1046 | hCoV-19/England/ALDP-C39372/2020\|EPI_ISL_731511\|20 | ···················· | ···················· | ···················· | ···················· | ···················· | ···················· |
| Y | 1047 | hCoV-19/England/ALDP-C3956D/2020\|EPI_ISL_731512\|20 | ···················· | ···················· | ···················· | ···················· | ···················· | ···················· |
| Y | 1048 | hCoV-19/England/QEUH-C31028/2020\|EPI_ISL_731548\|20 | ···················· | ···················· | ···················· | ···················· | ···················· | ···················· |
| Y | 1049 | hCoV-19/England/ALDP-C3A60F/2020\|EPI_ISL_731549\|20 | ···················· | ···················· | ···················· | ···················· | ···················· | ···················· |
| Y | 1057 | hCoV-19/Spain/RI-IBV-99010980/2020\|EPI_ISL_732807\| | ···················· | ···················· | ···················· | ···················· | ···················· | ···················· |
| Y | 1066 | hCoV-19/Spain/RI-IBV-99010993/2020\|EPI_ISL_732816\| | ···················· | ···················· | ···················· | ···················· | ···················· | ···················· |
| Y | 1070 | hCoV-19/USA/NE-001-17/2020\|EPI_ISL_732820\|2020-09- | ···················· | ···················· | ···················· | ···················· | ···················· | ···················· |
| Y | 1071 | hCoV-19/USA/NE-002-17/2020\|EPI_ISL_732821\|2020-09- | ···················· | ···················· | ···················· | ···················· | ···················· | ···················· |
| Y | 1072 | hCoV-19/USA/NE-003-17/2020\|EPI_ISL_732822\|2020-09- | ···················· | ···················· | ···················· | ···················· | ···················· | ···················· |

| | | | | | | | |
|---|---|---|---|---|---|---|---|
| Y | 1073 hCoV-19/USA/NE-004-17/2020\|EPI_ISL_732823\|2020-09- | ···················· | ···················· | ····················· | ···················· | ···················· | ···················· |
| Y | 1074 hCoV-19/USA/NE-005-17/2020\|EPI_ISL_732824\|2020-09- | ···················· | ···················· | ····················· | ···················· | ···················· | ···················· |
| Y | 1075 hCoV-19/USA/NE-006-17/2020\|EPI_ISL_732825\|2020-09- | ···················· | ···················· | ····················· | ···················· | ···················· | ···················· |
| Y | 1076 hCoV-19/USA/NE-007-17/2020\|EPI_ISL_732826\|2020-09- | ···················· | ···················· | ····················· | ···················· | ···················· | ···················· |
| Y | 1077 hCoV-19/Poland/77136/2020\|EPI_ISL_732828\|2020-11 | ···················· | ···················· | ····················· | ···················· | ···················· | ···················· |
| Y | 1078 hCoV-19/Poland/76226/2020\|EPI_ISL_732829\|2020-11 | ···················· | ···················· | ····················· | ···················· | ···················· | ···················· |
| Y | 1079 hCoV-19/Poland/76004/2020\|EPI_ISL_732830\|2020-11 | ···················· | ···················· | ····················· | ···················· | ···················· | ···················· |
| Y | 1080 hCoV-19/Poland/76343/2020\|EPI_ISL_732831\|2020-11 | ···················· | ···················· | ····················· | ···················· | ···················· | ···················· |
| Y | 1081 hCoV-19/Poland/77052/2020\|EPI_ISL_732832\|2020-11 | ···················· | ···················· | ····················· | ···················· | ···················· | ···················· |
| Y | 1082 hCoV-19/Poland/77173/2020\|EPI_ISL_732833\|2020-11 | ···················· | ···················· | ····················· | ···················· | ···················· | ···················· |
| Y | 1083 hCoV-19/Poland/76085/2020\|EPI_ISL_732834\|2020-11 | ···················· | ···················· | ····················· | ···················· | ···················· | ···················· |
| Y | 1084 hCoV-19/Poland/76311/2020\|EPI_ISL_732835\|2020-11 | ···················· | ···················· | ····················· | ···················· | ···················· | ···················· |
| Y | 1085 hCoV-19/Poland/76092/2020\|EPI_ISL_732836\|2020-11 | ···················· | ···················· | ····················· | ···················· | ···················· | ···················· |
| Y | 1086 hCoV-19/Poland/76133/2020\|EPI_ISL_732837\|2020-11 | ···················· | ···················· | ····················· | ···················· | ···················· | ···················· |
| Y | 1087 hCoV-19/Poland/76313/2020\|EPI_ISL_732838\|2020-11 | ···················· | ···················· | ····················· | ···················· | ···················· | ···················· |
| Y | 1088 hCoV-19/Poland/76703/2020\|EPI_ISL_732839\|2020-11 | ···················· | ···················· | ····················· | ···················· | ···················· | ···················· |
| Y | 1089 hCoV-19/Poland/77234/2020\|EPI_ISL_732840\|2020-11 | ···················· | ···················· | ····················· | ···················· | ···················· | ···················· |
| Y | 1090 hCoV-19/Poland/76002/2020\|EPI_ISL_732841\|2020-11 | ········T··········· | ···················· | ····················· | ···················· | ···················· | ···················· |
| Y | 1091 hCoV-19/Poland/76144/2020\|EPI_ISL_732842\|2020-11 | ···················· | ···················· | ····················· | ···················· | ···················· | ···················· |
| Y | 1092 hCoV-19/Poland/77220/2020\|EPI_ISL_732843\|2020-11 | ···················· | ···················· | ····················· | ···················· | ···················· | ···················· |
| Y | 1093 hCoV-19/Poland/76220/2020\|EPI_ISL_732879\|2020-11 | ···················· | ···················· | ····················· | ···················· | ···················· | ···················· |
| Y | 1094 hCoV-19/Belgium/rega-0918996/2020\|EPI_ISL_735000\|2 | ···················· | ···················· | ····················· | ···················· | ···················· | ···················· |
| Y | 1095 hCoV-19/Belgium/rega-09191058/2020\|EPI_ISL_735001\| | ···················· | ···················· | ····················· | ···················· | ···················· | ···················· |
| Y | 1096 hCoV-19/Belgium/rega-09191059/2020\|EPI_ISL_735002\| | ···················· | ···················· | ····················· | ···················· | ···················· | ···················· |
| Y | 1097 hCoV-19/Belgium/rega-09191060/2020\|EPI_ISL_735003\| | ···················· | ···················· | ····················· | ···················· | ···················· | ···················· |
| Y | 1098 hCoV-19/Belgium/rega-09201062/2020\|EPI_ISL_735004\| | ···················· | ···················· | ····················· | ···················· | ···················· | ···················· |
| Y | 1099 hCoV-19/Belgium/rega-09201063/2020\|EPI_ISL_735005\| | ···················· | ···················· | ····················· | ···················· | ···················· | ···················· |
| Y | 1100 hCoV-19/Belgium/rega-09211061/2020\|EPI_ISL_735006\| | ···················· | ···················· | ····················· | ···················· | ···················· | ···················· |
| Y | 1101 hCoV-19/Belgium/rega-09221000/2020\|EPI_ISL_735007\| | ···················· | ···················· | ····················· | ···················· | ···················· | ···················· |
| Y | 1102 hCoV-19/Belgium/rega-09221001/2020\|EPI_ISL_735008\| | ···················· | ···················· | ····················· | ···················· | ···················· | ···················· |
| Y | 1103 hCoV-19/Belgium/rega-09221002/2020\|EPI_ISL_735009\| | ···················· | ···················· | ····················· | ···················· | ···················· | ···················· |
| Y | 1104 hCoV-19/Belgium/rega-09221012/2020\|EPI_ISL_735010\| | ···················· | ···················· | ····················· | ···················· | ···················· | ···················· |
| Y | 1105 hCoV-19/Belgium/rega-09221013/2020\|EPI_ISL_735011\| | ···················· | ···················· | ····················· | ···················· | ···················· | ···················· |
| Y | 1106 hCoV-19/Belgium/rega-09221014/2020\|EPI_ISL_735012\| | ···················· | ···················· | ····················· | ···················· | ···················· | ···················· |
| Y | 1107 hCoV-19/Belgium/rega-09221015/2020\|EPI_ISL_735013\| | ···················· | ···················· | ····················· | ···················· | ···················· | ···················· |
| Y | 1108 hCoV-19/Belgium/rega-09221016/2020\|EPI_ISL_735014\| | ···················· | ···················· | ····················· | ···················· | ···················· | ···················· |
| Y | 1109 hCoV-19/Belgium/rega-09221017/2020\|EPI_ISL_735015\| | ···················· | ···················· | ····················· | ···················· | ···················· | ···················· |
| Y | 1110 hCoV-19/Belgium/rega-09221018/2020\|EPI_ISL_735016\| | ···················· | ···················· | ····················· | ···················· | ···················· | ···················· |
| Y | 1111 hCoV-19/Belgium/rega-09221019/2020\|EPI_ISL_735017\| | ···················· | ···················· | ····················· | ···················· | ···················· | ···················· |
| Y | 1112 hCoV-19/Belgium/rega-09221020/2020\|EPI_ISL_735018\| | ···················· | ···················· | ····················· | ···················· | ···················· | ···················· |
| Y | 1113 hCoV-19/Belgium/rega-09221021/2020\|EPI_ISL_735019\| | ···················· | ···················· | ····················· | ···················· | ···················· | ···················· |
| Y | 1114 hCoV-19/Belgium/rega-09221022/2020\|EPI_ISL_735020\| | ···················· | ···················· | ····················· | ···················· | ···················· | ···················· |
| Y | 1115 hCoV-19/Belgium/rega-09241236/2020\|EPI_ISL_735040\| | ···················· | ···················· | ····················· | ···················· | ···················· | ···················· |
| Y | 1116 hCoV-19/Belgium/rega-09241241/2020\|EPI_ISL_735041\| | ···················· | ···················· | ····················· | ···················· | ···················· | ···················· |
| Y | 1117 hCoV-19/Belgium/rega-09241242/2020\|EPI_ISL_735042\| | ···················· | ···················· | ····················· | ···················· | ······T············· | ···················· |
| Y | 1118 hCoV-19/Belgium/rega-09241253/2020\|EPI_ISL_735043\| | ···················· | ···················· | ····················· | ···················· | ······T············· | ···················· |
| Y | 1119 hCoV-19/Belgium/rega-10031026/2020\|EPI_ISL_735044\| | ···················· | ···················· | ····················· | ···················· | ···················· | ···················· |
| Y | 1120 hCoV-19/Belgium/rega-10031027/2020\|EPI_ISL_735045\| | ···················· | ···················· | ····················· | ···················· | ···················· | ···················· |
| Y | 1121 hCoV-19/Belgium/rega-10031028/2020\|EPI_ISL_735046\| | ···················· | ···················· | ····················· | ···················· | ···················· | ···················· |
| Y | 1122 hCoV-19/Belgium/rega-10031030/2020\|EPI_ISL_735047\| | ···················· | ···················· | ····················· | ···················· | ···················· | ···················· |
| Y | 1123 hCoV-19/Belgium/rega-10031031/2020\|EPI_ISL_735048\| | ···················· | ···················· | ····················· | ···················· | ···················· | ···················· |
| Y | 1124 hCoV-19/Belgium/rega-10061009/2020\|EPI_ISL_735049\| | ···················· | ···················· | ····················· | ···················· | ···················· | ···················· |
| Y | 1125 hCoV-19/Belgium/rega-10061010/2020\|EPI_ISL_735050\| | ···················· | ···················· | ····················· | ···················· | ···················· | ···················· |
| Y | 1126 hCoV-19/Belgium/rega-10061011/2020\|EPI_ISL_735051\| | ···················· | ···················· | ····················· | ···················· | ······T············· | ···················· |
| Y | 1127 hCoV-19/Belgium/rega-10061029/2020\|EPI_ISL_735052\| | ···················· | ···················· | ····················· | ···················· | ···················· | ···················· |
| Y | 1128 hCoV-19/Belgium/rega-10061032/2020\|EPI_ISL_735053\| | ···················· | ···················· | ····················· | ···················· | ···················· | ···················· |
| Y | 1129 hCoV-19/Belgium/rega-10061033/2020\|EPI_ISL_735054\| | ···················· | ···················· | ····················· | ···················· | ···················· | ···················· |
| Y | 1130 hCoV-19/Belgium/rega-10061034/2020\|EPI_ISL_735055\| | ···················· | ···················· | ····················· | ···················· | ···················· | ···················· |
| Y | 1131 hCoV-19/Belgium/rega-10061035/2020\|EPI_ISL_735056\| | ···················· | ···················· | ····················· | ···················· | ···················· | ···················· |
| Y | 1132 hCoV-19/Belgium/rega-10061037/2020\|EPI_ISL_735057\| | ···················· | ···················· | ····················· | ···················· | ···················· | ···················· |
| Y | 1133 hCoV-19/Belgium/rega-10061038/2020\|EPI_ISL_735058\| | ···················· | ···················· | ····················· | ···················· | ···················· | ···················· |
| Y | 1134 hCoV-19/Belgium/rega-10061039/2020\|EPI_ISL_735059\| | ···················· | ···················· | ····················· | ···················· | ···················· | ···················· |
| Y | 1135 hCoV-19/Belgium/rega-10061040/2020\|EPI_ISL_735060\| | ···················· | ···················· | ····················· | ···················· | ···················· | ···················· |
| Y | 1136 hCoV-19/Belgium/rega-10061041/2020\|EPI_ISL_735061\| | ···················· | ···················· | ····················· | ···················· | ···················· | ···················· |
| Y | 1137 hCoV-19/Belgium/rega-10061042/2020\|EPI_ISL_735062\| | ···················· | ····A··············· | ····················· | ···················· | ···················· | ···················· |
| Y | 1138 hCoV-19/Belgium/rega-10061043/2020\|EPI_ISL_735063\| | ···················· | ···················· | ····················· | ···················· | ···················· | ···················· |
| Y | 1139 hCoV-19/Belgium/rega-10061044/2020\|EPI_ISL_735064\| | ···················· | ···················· | ····················· | ···················· | ···················· | ···················· |
| Y | 1140 hCoV-19/Belgium/rega-10061045/2020\|EPI_ISL_735065\| | ···················· | ···················· | ····················· | ···················· | ···················· | ···················· |
| Y | 1141 hCoV-19/Belgium/rega-10061046/2020\|EPI_ISL_735066\| | ···················· | ···················· | ····················· | ···················· | ···················· | ···················· |
| Y | 1142 hCoV-19/Belgium/rega-10061193/2020\|EPI_ISL_735067\| | ···················· | ···················· | ····················· | ···················· | ···················· | ···················· |
| Y | 1143 hCoV-19/Belgium/rega-10061194/2020\|EPI_ISL_735068\| | ···················· | ···················· | ····················· | ···················· | ···················· | ···················· |
| Y | 1144 hCoV-19/Belgium/rega-10061195/2020\|EPI_ISL_735069\| | ···················· | ···················· | ····················· | ···················· | ···················· | ···················· |
| Y | 1145 hCoV-19/Belgium/rega-10061196/2020\|EPI_ISL_735070\| | ···················· | ···················· | ····················· | ···················· | ···················· | ···················· |
| Y | 1146 hCoV-19/Belgium/rega-10061197/2020\|EPI_ISL_735071\| | ···················· | ···················· | ····················· | ···················· | ···················· | ···················· |
| Y | 1147 hCoV-19/Belgium/rega-10061198/2020\|EPI_ISL_735072\| | ···················· | ···················· | ····················· | ···················· | ···················· | ···················· |
| Y | 1148 hCoV-19/Belgium/rega-10061213/2020\|EPI_ISL_735073\| | ···················· | ···················· | ····················· | ···················· | ···················· | ···················· |
| Y | 1149 hCoV-19/Belgium/rega-10061255/2020\|EPI_ISL_735074\| | ···················· | ···················· | ····················· | ···················· | ···················· | ···················· |
| Y | 1150 hCoV-19/Belgium/rega-10071231/2020\|EPI_ISL_735075\| | ···················· | ···················· | ····················· | ···················· | ···················· | ···················· |
| Y | 1151 hCoV-19/Belgium/rega-10071232/2020\|EPI_ISL_735076\| | ···················· | ···················· | ····················· | ···················· | ···················· | ···················· |
| Y | 1152 hCoV-19/Belgium/rega-10071237/2020\|EPI_ISL_735077\| | ···················· | ···················· | ····················· | ···················· | ···················· | ···················· |

| | | | | | | | |
|---|---|---|---|---|---|---|---|
| Y | 1153 hCoV-19/Belgium/rega-10071243/2020\|EPI_ISL_735078\| | ···················· | ···················· | ····················· | ···················· | ···················· | ···················· |
| Y | 1154 hCoV-19/Belgium/rega-10071249/2020\|EPI_ISL_735079\| | ···················· | ···················· | ····················· | ···················· | ···················· | ···················· |
| Y | 1155 hCoV-19/Belgium/rega-10081214/2020\|EPI_ISL_735080\| | ···················· | ···················· | ····················· | ···················· | ···················· | ···················· |
| Y | 1156 hCoV-19/Belgium/rega-10081215/2020\|EPI_ISL_735081\| | ···················· | ···················· | ····················· | ···················· | ···················· | ···················· |
| Y | 1157 hCoV-19/Belgium/rega-10081220/2020\|EPI_ISL_735082\| | ···················· | ···················· | ····················· | ···················· | ···················· | ···················· |
| Y | 1158 hCoV-19/Belgium/rega-10081221/2020\|EPI_ISL_735083\| | ···················· | ···················· | ····················· | ···················· | ···················· | ···················· |
| Y | 1159 hCoV-19/Belgium/rega-10081226/2020\|EPI_ISL_735084\| | ···················· | ···················· | ····················· | ···················· | ···················· | ···················· |
| Y | 1160 hCoV-19/Belgium/rega-10081227/2020\|EPI_ISL_735085\| | ···················· | ···················· | ····················· | ···················· | ···················· | ···················· |
| Y | 1161 hCoV-19/Belgium/rega-10081233/2020\|EPI_ISL_735086\| | ···················· | ···················· | ····················· | ···················· | ···················· | ···················· |
| Y | 1162 hCoV-19/Belgium/rega-10081238/2020\|EPI_ISL_735087\| | ···················· | ···················· | ····················· | ···················· | ···················· | ···················· |
| Y | 1163 hCoV-19/Belgium/rega-10081239/2020\|EPI_ISL_735088\| | ···················· | ···················· | ····················· | ···················· | ···················· | ···················· |
| Y | 1164 hCoV-19/Belgium/rega-10081244/2020\|EPI_ISL_735089\| | ···················· | ···················· | ····················· | ···················· | ···················· | ···················· |
| Y | 1165 hCoV-19/Belgium/rega-10081245/2020\|EPI_ISL_735090\| | ···················· | ···················· | ····················· | ···················· | ···················· | ···················· |
| Y | 1166 hCoV-19/Belgium/rega-10081250/2020\|EPI_ISL_735091\| | ···················· | ···················· | ····················· | ···················· | ···················· | ···················· |
| Y | 1167 hCoV-19/Belgium/rega-10081251/2020\|EPI_ISL_735092\| | ···················· | ···················· | ····················· | ···················· | ···················· | ···················· |
| Y | 1168 hCoV-19/Belgium/rega-10081256/2020\|EPI_ISL_735093\| | ···················· | ···················· | ····················· | ···················· | ···················· | ···················· |
| Y | 1170 hCoV-19/Belgium/rega-10151114/2020\|EPI_ISL_735095\| | ···················· | ···················· | ····················· | ···················· | ···················· | ···················· |
| Y | 1171 hCoV-19/Belgium/rega-10151115/2020\|EPI_ISL_735096\| | ···················· | ···················· | ····················· | ···················· | ···················· | ···················· |
| Y | 1172 hCoV-19/Belgium/rega-10151116/2020\|EPI_ISL_735097\| | ···················· | ···················· | ····················· | ···················· | ···················· | ···················· |
| Y | 1173 hCoV-19/Belgium/rega-10151118/2020\|EPI_ISL_735098\| | ···················· | ···················· | ····················· | ···················· | ···················· | ···················· |
| Y | 1174 hCoV-19/Belgium/rega-10151119/2020\|EPI_ISL_735099\| | ···················· | ···················· | ····················· | ···················· | ···················· | ···················· |
| Y | 1175 hCoV-19/England/QEUH-C579A9/2020\|EPI_ISL_736330\|20 | ···················· | ···················· | ····················· | ···················· | ···················· | ···················· |
| Y | 1176 hCoV-19/England/QEUH-C579C7/2020\|EPI_ISL_736331\|20 | ···················· | ···················· | ····················· | ···················· | ···················· | ···················· |
| Y | 1177 hCoV-19/England/QEUH-C579B8/2020\|EPI_ISL_736332\|20 | ···················· | ···················· | ····················· | ···················· | ···················· | ···················· |
| Y | 1178 hCoV-19/England/MILK-C52E27/2020\|EPI_ISL_736333\|20 | ···················· | ···················· | ····················· | ···················· | ···················· | ···················· |
| Y | 1179 hCoV-19/England/QEUH-C53E9F/2020\|EPI_ISL_736334\|20 | ···················· | ···················· | ····················· | ···················· | ···················· | ···················· |
| Y | 1180 hCoV-19/England/MILK-C53CE0/2020\|EPI_ISL_736342\|20 | ···················· | ···················· | ····················· | ···················· | ···················· | ···················· |
| Y | 1181 hCoV-19/England/QEUH-C54098/2020\|EPI_ISL_736347\|20 | ···················· | ···················· | ····················· | ···················· | ···················· | ···················· |
| Y | 1182 hCoV-19/England/QEUH-C54669/2020\|EPI_ISL_736370\|20 | ···················· | ···················· | ····················· | ···················· | ···················· | ···················· |
| Y | 1183 hCoV-19/England/QEUH-C57A88/2020\|EPI_ISL_736371\|20 | ···················· | ···················· | ····················· | ···················· | ···················· | ···················· |
| Y | 1184 hCoV-19/England/QEUH-C57BB2/2020\|EPI_ISL_736373\|20 | ···················· | ···················· | ····················· | ···················· | ···················· | ···················· |
| Y | 1185 hCoV-19/England/MILK-C52D2A/2020\|EPI_ISL_736374\|20 | ···················· | ···················· | ····················· | ···················· | ···················· | ···················· |
| Y | 1186 hCoV-19/England/QEUH-C5470C/2020\|EPI_ISL_736376\|20 | ···················· | ···················· | ····················· | ···················· | ···················· | ···················· |
| Y | 1187 hCoV-19/England/QEUH-C53F50/2020\|EPI_ISL_736378\|20 | ···················· | ···················· | ····················· | ···················· | ···················· | ···················· |
| Y | 1188 hCoV-19/England/MILK-C52C1E/2020\|EPI_ISL_736385\|20 | ···················· | ···················· | ····················· | ···················· | ···················· | ···················· |
| Y | 1189 hCoV-19/England/MILK-C52D48/2020\|EPI_ISL_736387\|20 | ···················· | ···················· | ····················· | ···················· | ···················· | ···················· |
| Y | 1190 hCoV-19/England/QEUH-C57A97/2020\|EPI_ISL_736388\|20 | ···················· | ···················· | ····················· | ···················· | ···················· | ···················· |
| Y | 1191 hCoV-19/England/QEUH-C54010/2020\|EPI_ISL_736389\|20 | ···················· | ···················· | ····················· | ···················· | ···················· | ···················· |
| Y | 1192 hCoV-19/England/QEUH-C53E08/2020\|EPI_ISL_736390\|20 | ···················· | ···················· | ····················· | ···················· | ···················· | ···················· |
| Y | 1193 hCoV-19/England/QEUH-C53FD8/2020\|EPI_ISL_736391\|20 | ···················· | ···················· | ····················· | ···················· | ···················· | ···················· |
| Y | 1194 hCoV-19/England/QEUH-C579E5/2020\|EPI_ISL_736394\|20 | ···················· | ···················· | ····················· | ···················· | ···················· | ···················· |
| Y | 1195 hCoV-19/England/QEUH-C54520/2020\|EPI_ISL_736398\|20 | ···················· | ···················· | ····················· | ···················· | ···················· | ···················· |
| Y | 1196 hCoV-19/Switzerland/AG-ETHZ-410960/2020\|EPI_ISL_73 | ···················· | ···················· | ····················· | ···················· | ···················· | ···················· |
| Y | 1197 hCoV-19/Switzerland/AR-ETHZ-410900/2020\|EPI_ISL_73 | ···················· | ···················· | ····················· | ···················· | ···················· | ···················· |
| Y | 1198 hCoV-19/Switzerland/BL-ETHZ-410054/2020\|EPI_ISL_73 | ···················· | ···················· | ····················· | ···················· | ···················· | ···················· |
| Y | 1199 hCoV-19/Switzerland/BL-ETHZ-410056/2020\|EPI_ISL_73 | ···················· | ···················· | ····················· | ···················· | ···················· | ···················· |
| Y | 1201 hCoV-19/Switzerland/BL-ETHZ-410141/2020\|EPI_ISL_73 | ···················· | ·A·················· | ····················· | ···················· | ···················· | ··A················· |
| Y | 1202 hCoV-19/Switzerland/BL-ETHZ-410175/2020\|EPI_ISL_73 | ···················· | ···················· | ····················· | ···················· | ···················· | ···················· |
| Y | 1203 hCoV-19/Switzerland/BL-ETHZ-410181/2020\|EPI_ISL_73 | ···················· | ···················· | ····················· | ···················· | ···················· | ···················· |
| Y | 1205 hCoV-19/Switzerland/BL-ETHZ-410193/2020\|EPI_ISL_73 | ···················· | ···················· | ····················· | ···················· | ···················· | ···················· |
| Y | 1206 hCoV-19/Switzerland/BL-ETHZ-410252/2020\|EPI_ISL_73 | ···················· | ···················· | ····················· | ···················· | ···················· | ···················· |
| Y | 1207 hCoV-19/Switzerland/BL-ETHZ-410254/2020\|EPI_ISL_73 | ···················· | ···················· | ····················· | ···················· | ···················· | ···················· |
| Y | 1208 hCoV-19/Switzerland/BL-ETHZ-410257/2020\|EPI_ISL_73 | ···················· | ···················· | ····················· | ···················· | ···················· | ···················· |
| Y | 1209 hCoV-19/Switzerland/BL-ETHZ-410265/2020\|EPI_ISL_73 | ···················· | ·········A·········· | ····················· | ···················· | ···················· | ···················· |
| Y | 1210 hCoV-19/Switzerland/BL-ETHZ-410277/2020\|EPI_ISL_73 | ···················· | ···················· | ····················· | ···················· | ···················· | ···················· |
| Y | 1211 hCoV-19/Switzerland/BL-ETHZ-410278/2020\|EPI_ISL_73 | ···················· | ···················· | ····················· | ···················· | ···················· | ···················· |
| Y | 1212 hCoV-19/Switzerland/BL-ETHZ-410818/2020\|EPI_ISL_73 | ···················· | ···················· | ····················· | ···················· | ···················· | ···················· |
| Y | 1213 hCoV-19/Switzerland/BL-ETHZ-410862/2020\|EPI_ISL_73 | ···················· | ···················· | ····················· | ···················· | ···················· | ···················· |
| Y | 1214 hCoV-19/Switzerland/BL-ETHZ-410863/2020\|EPI_ISL_73 | ···················· | ···················· | ····················· | ···················· | ···················· | ···················· |
| Y | 1215 hCoV-19/Switzerland/BL-ETHZ-410953/2020\|EPI_ISL_73 | ···················· | ···················· | ····················· | ···················· | ···················· | ···················· |
| Y | 1217 hCoV-19/USA/MT-CDC-2-3693670/2020\|EPI_ISL_747032\|2 | ···················· | ···················· | ····················· | ···················· | ···················· | ···················· |
| Y | 1218 hCoV-19/USA/MT-CDC-2-3693754/2020\|EPI_ISL_747033\|2 | ···················· | ···················· | ····················· | ···················· | ···················· | ···················· |
| Y | 1219 hCoV-19/USA/MT-CDC-2-3693496/2020\|EPI_ISL_747034\|2 | ···················· | ···················· | ····················· | ···················· | ···················· | ···················· |
| Y | 1220 hCoV-19/USA/MT-CDC-2-3693787/2020\|EPI_ISL_747035\|2 | ···················· | ·A·················· | ····················· | ···················· | ···················· | ···················· |
| Y | 1221 hCoV-19/USA/DE-CDC-2-3693673/2020\|EPI_ISL_747036\|2 | ···················· | ·A·················· | ····················· | ···················· | ···················· | ···················· |
| Y | 1222 hCoV-19/USA/DE-CDC-2-3693785/2020\|EPI_ISL_747038\|2 | ···················· | ·A·················· | ····················· | ···················· | ···················· | ···················· |
| Y | 1223 hCoV-19/USA/NY-CDC-2-3693757/2020\|EPI_ISL_747039\|2 | ···················· | ···················· | ····················· | ···················· | ···················· | ···················· |
| Y | 1224 hCoV-19/USA/NY-CDC-2-3693492/2020\|EPI_ISL_747040\|2 | ···················· | ···················· | ····················· | ···················· | ···················· | ···················· |
| Y | 1225 hCoV-19/USA/NY-CDC-2-3693655/2020\|EPI_ISL_747041\|2 | ···················· | ·A·················· | ····················· | ···················· | ···················· | ···················· |
| Y | 1226 hCoV-19/USA/NY-CDC-2-3693796/2020\|EPI_ISL_747042\|2 | ···················· | ···················· | ····················· | ···················· | ···················· | ···················· |
| Y | 1227 hCoV-19/USA/NY-CDC-2-3693767/2020\|EPI_ISL_747043\|2 | ···················· | ·A·················· | ····················· | ···················· | ···················· | ···················· |
| Y | 1228 hCoV-19/USA/AL-CDC-2-3693782/2020\|EPI_ISL_747044\|2 | ···················· | ···················· | ····················· | ···················· | ···················· | ···················· |
| Y | 1229 hCoV-19/USA/AL-CDC-2-3693696/2020\|EPI_ISL_747046\|2 | ···················· | ···················· | ····················· | ···················· | ···················· | ···················· |
| Y | 1230 hCoV-19/USA/AL-CDC-2-3693697/2020\|EPI_ISL_747047\|2 | ···················· | ···················· | ····················· | ···················· | ···················· | ···················· |
| Y | 1231 hCoV-19/USA/AL-CDC-2-3693671/2020\|EPI_ISL_747048\|2 | ···················· | ···················· | ····················· | ···················· | ···················· | ···················· |
| Y | 1232 hCoV-19/USA/PA-CDC-2-3693775/2020\|EPI_ISL_747049\|2 | ···················· | ···················· | ····················· | ···················· | ···················· | ···················· |
| Y | 1233 hCoV-19/USA/PA-CDC-2-3693664/2020\|EPI_ISL_747050\|2 | ···················· | ·A·················· | ····················· | ···················· | ···················· | ···················· |
| Y | 1234 hCoV-19/USA/PA-CDC-2-3693774/2020\|EPI_ISL_747051\|2 | ···················· | ···················· | ····················· | ···················· | ···················· | ···················· |
| Y | 1235 hCoV-19/USA/PA-CDC-2-3693665/2020\|EPI_ISL_747052\|2 | ···················· | ···················· | ····················· | ···················· | ···················· | ···················· |
| Y | 1236 hCoV-19/USA/PA-CDC-2-3693751/2020\|EPI_ISL_747053\|2 | ···················· | ···················· | ····················· | ···················· | ···················· | ···················· |

| | | | | | | | |
|---|---|---|---|---|---|---|---|
| Y | 1237 hCoV-19/USA/AK-CDC-2-3693667/2020\|EPI_ISL_747054\|2 | .................... | .................... | .................... | .................... | .................... | .................... |
| Y | 1238 hCoV-19/USA/AK-CDC-2-3693690/2020\|EPI_ISL_747055\|2 | .................... | .................... | .................... | .................... | .................... | .................... |
| Y | 1239 hCoV-19/USA/AK-CDC-2-3693795/2020\|EPI_ISL_747056\|2 | .................... | .................... | .................... | .................... | .................... | .................... |
| Y | 1240 hCoV-19/USA/AK-CDC-2-3693677/2020\|EPI_ISL_747057\|2 | .................... | .................... | .................... | .................... | .................... | .................... |
| Y | 1241 hCoV-19/USA/AK-CDC-2-3693693/2020\|EPI_ISL_747058\|2 | .................... | .................... | .................... | .................... | .................... | .................... |
| Y | 1242 hCoV-19/USA/TN-CDC-2-3693791/2020\|EPI_ISL_747059\|2 | .................... | .............A...... | .................... | .................... | .................... | .................... |
| Y | 1243 hCoV-19/USA/TN-CDC-2-3693694/2020\|EPI_ISL_747061\|2 | .................... | .................... | .................... | .................... | .................... | .................... |
| Y | 1244 hCoV-19/USA/TN-CDC-2-3693660/2020\|EPI_ISL_747062\|2 | .................... | .................... | .................... | .................... | .................... | .................... |
| Y | 1245 hCoV-19/USA/TN-CDC-2-3693653/2020\|EPI_ISL_747063\|2 | .................... | .................... | .................... | .................... | .................... | .................... |
| Y | 1246 hCoV-19/USA/AR-CDC-2-3693765/2020\|EPI_ISL_747064\|2 | .................... | .................... | .................... | .................... | .................... | .................... |
| Y | 1247 hCoV-19/USA/AR-CDC-2-3693682/2020\|EPI_ISL_747065\|2 | .................... | .................... | .................... | .................... | .................... | .................... |
| Y | 1248 hCoV-19/USA/AR-CDC-2-3693681/2020\|EPI_ISL_747066\|2 | .................... | .................... | .................... | .................... | .................... | .................... |
| Y | 1249 hCoV-19/USA/AR-CDC-2-3693691/2020\|EPI_ISL_747067\|2 | .................... | .A.................. | .................... | .................... | .................... | .................... |
| Y | 1250 hCoV-19/USA/AR-CDC-2-3693771/2020\|EPI_ISL_747068\|2 | .................... | .................... | .................... | .................... | .................... | .................... |
| Y | 1251 hCoV-19/USA/LA-CDC-2-3693793/2020\|EPI_ISL_747069\|2 | .................... | .................... | .................... | .................... | .................... | .................... |
| Y | 1252 hCoV-19/USA/LA-CDC-2-3693763/2020\|EPI_ISL_747070\|2 | .................... | .................... | .................... | .................... | .................... | .................... |
| Y | 1253 hCoV-19/USA/LA-CDC-2-3693778/2020\|EPI_ISL_747071\|2 | .................... | .................... | .................... | .................... | .................... | .................... |
| Y | 1254 hCoV-19/USA/LA-CDC-2-3693761/2020\|EPI_ISL_747072\|2 | .................... | .A.................. | .................... | .................... | .................... | .................... |
| Y | 1255 hCoV-19/USA/LA-CDC-2-3693755/2020\|EPI_ISL_747073\|2 | .................... | .A.................. | .................... | .................... | .................... | .................... |
| Y | 1256 hCoV-19/USA/UT-CDC-2-3693760/2020\|EPI_ISL_747074\|2 | .................... | .................... | .................... | .................... | .................... | .................... |
| Y | 1257 hCoV-19/USA/UT-CDC-2-3693769/2020\|EPI_ISL_747075\|2 | .................... | .................... | .................... | .................... | .................... | .................... |
| Y | 1258 hCoV-19/USA/UT-CDC-2-3693656/2020\|EPI_ISL_747076\|2 | .................... | .................... | .................... | .................... | .................... | .................... |
| Y | 1259 hCoV-19/USA/UT-CDC-2-3693661/2020\|EPI_ISL_747077\|2 | .................... | .................... | .................... | .................... | .................... | .................... |
| Y | 1260 hCoV-19/USA/WI-CDC-2-3693753/2020\|EPI_ISL_747079\|2 | .................... | .................... | .................... | .................... | .................... | .................... |
| Y | 1261 hCoV-19/USA/WI-CDC-2-3693678/2020\|EPI_ISL_747080\|2 | .................... | .................... | .................... | .................... | .................... | .................... |
| Y | 1262 hCoV-19/USA/WI-CDC-2-3693658/2020\|EPI_ISL_747081\|2 | .................... | .................... | .................... | .................... | .................... | .................... |
| Y | 1263 hCoV-19/USA/WI-CDC-2-3693669/2020\|EPI_ISL_747082\|2 | .................... | .................... | .................... | .................... | .................... | .................... |
| Y | 1264 hCoV-19/USA/WI-CDC-2-3693692/2020\|EPI_ISL_747083\|2 | .................... | .................... | .................... | .................... | T................... | .................... |
| Y | 1265 hCoV-19/USA/IN-CDC-2-3693790/2020\|EPI_ISL_747084\|2 | .................... | .................... | .................... | .................... | .................... | .................... |
| Y | 1266 hCoV-19/USA/IN-CDC-2-3693679/2020\|EPI_ISL_747085\|2 | .................... | .................... | .................... | .................... | .................... | .................... |
| Y | 1267 hCoV-19/USA/IN-CDC-2-3693675/2020\|EPI_ISL_747086\|2 | .................... | .................... | .................... | .................... | .................... | .................... |
| Y | 1268 hCoV-19/USA/IN-CDC-2-3693766/2020\|EPI_ISL_747087\|2 | .................... | .................... | .................... | .................... | .................... | .................... |
| Y | 1269 hCoV-19/USA/IN-CDC-2-3693654/2020\|EPI_ISL_747088\|2 | .................... | .................... | .................... | .................... | .................... | .................... |
| Y | 1270 hCoV-19/USA/KY-CDC-2-3693493/2020\|EPI_ISL_747089\|2 | .................... | .................... | .................... | .................... | .................... | .................... |
| Y | 1271 hCoV-19/USA/KY-CDC-2-3693781/2020\|EPI_ISL_747090\|2 | .................... | .A.................. | .................... | .................... | .................... | .................... |
| Y | 1272 hCoV-19/USA/KY-CDC-2-3693686/2020\|EPI_ISL_747091\|2 | .................... | .................... | .................... | .................... | .................... | .................... |
| Y | 1273 hCoV-19/USA/KY-CDC-2-3693794/2020\|EPI_ISL_747092\|2 | .................... | .................... | .................... | .................... | .................... | .................... |
| Y | 1274 hCoV-19/USA/KY-CDC-2-3693772/2020\|EPI_ISL_747093\|2 | .................... | .................... | .................... | .................... | .................... | .................... |
| Y | 1275 hCoV-19/USA/OK-CDC-2-3693792/2020\|EPI_ISL_747094\|2 | .................... | .................... | .................... | .................... | .................... | .................... |
| Y | 1276 hCoV-19/USA/OK-CDC-2-3693683/2020\|EPI_ISL_747095\|2 | .................... | .A.................. | .................... | .................... | .................... | .................... |
| Y | 1277 hCoV-19/USA/OK-CDC-2-3693756/2020\|EPI_ISL_747096\|2 | .................... | .A.................. | .................... | .................... | .................... | .................... |
| Y | 1278 hCoV-19/USA/OK-CDC-2-3693789/2020\|EPI_ISL_747097\|2 | .................... | .................... | .................... | .................... | .................... | .................... |
| Y | 1279 hCoV-19/USA/OK-CDC-2-3693672/2020\|EPI_ISL_747098\|2 | .................... | .A.................. | .................... | .................... | .................... | .................... |
| Y | 1280 hCoV-19/USA/MS-CDC-2-3693773/2020\|EPI_ISL_747099\|2 | .................... | .............A...... | .................... | .................... | .................... | .................... |
| Y | 1281 hCoV-19/Wales/PHWC-4927BF/2020\|EPI_ISL_725700\|2020 | .................... | .................... | .................... | .................... | .................... | .................... |
| Y | 1282 hCoV-19/Wales/PHWC-4927CE/2020\|EPI_ISL_725701\|2020 | .................... | .................... | .................... | .................... | .................... | .................... |
| Y | 1283 hCoV-19/Wales/PHWC-4927FB/2020\|EPI_ISL_725703\|2020 | .................... | .................... | .................... | .................... | .................... | .................... |
| Y | 1284 hCoV-19/Wales/PHWC-492816/2020\|EPI_ISL_725705\|2020 | .................... | .................... | .................... | .................... | .................... | .................... |
| Y | 1286 hCoV-19/Wales/PHWC-492843/2020\|EPI_ISL_725708\|2020 | .................... | .................... | .................... | .................... | .................... | .................... |
| Y | 1287 hCoV-19/Wales/PHWC-492852/2020\|EPI_ISL_725709\|2020 | .................... | .................... | .................... | .................... | .................... | .................... |
| Y | 1289 hCoV-19/Wales/PHWC-492870/2020\|EPI_ISL_725711\|2020 | .................... | .................... | .................... | .................... | .................... | .................... |
| Y | 1290 hCoV-19/Wales/PHWC-49289E/2020\|EPI_ISL_725713\|2020 | .................... | .................... | .................... | .................... | .................... | .................... |
| Y | 1292 hCoV-19/Wales/PHWC-4928CB/2020\|EPI_ISL_725715\|2020 | .................... | .................... | .................... | .................... | .................... | .................... |
| Y | 1293 hCoV-19/Switzerland/BL-ETHZ-350355/2020\|EPI_ISL_72 | .................... | .................... | .................... | .................... | .................... | .................... |
| Y | 1294 hCoV-19/Switzerland/GE-ETHZ-350434/2020\|EPI_ISL_72 | .................... | .................... | .................... | .................... | .................... | .................... |
| Y | 1295 hCoV-19/Switzerland/NE-ETHZ-350442/2020\|EPI_ISL_72 | .................... | .................... | .................... | .................... | .................... | .................... |
| Y | 1296 hCoV-19/Switzerland/AG-ETHZ-350447/2020\|EPI_ISL_72 | .................... | .................... | .................... | .................... | .................... | .................... |
| Y | 1297 hCoV-19/Switzerland/NE-ETHZ-350456/2020\|EPI_ISL_72 | .................... | .................... | .................... | .................... | .................... | .................... |
| Y | 1298 hCoV-19/Switzerland/NE-ETHZ-350457/2020\|EPI_ISL_72 | .................... | .................... | .................... | .................... | .................... | .................... |
| Y | 1299 hCoV-19/Switzerland/ZH-ETHZ-350458/2020\|EPI_ISL_72 | .................... | .................... | .................... | .................... | .................... | .................... |
| Y | 1300 hCoV-19/Switzerland/NE-ETHZ-350460/2020\|EPI_ISL_72 | .................... | .................... | .................... | .................... | .................... | .................... |
| Y | 1301 hCoV-19/Switzerland/NE-ETHZ-350463/2020\|EPI_ISL_72 | .................... | .................... | .................... | .................... | .................... | .................... |
| Y | 1302 hCoV-19/Switzerland/BE-ETHZ-350471/2020\|EPI_ISL_72 | .................... | .................... | .................... | .................... | .................... | .................... |
| Y | 1303 hCoV-19/Switzerland/GR-ETHZ-350476/2020\|EPI_ISL_72 | .................... | .................... | .................... | .................... | .................... | .................... |
| Y | 1304 hCoV-19/Switzerland/BE-ETHZ-360343/2020\|EPI_ISL_72 | .................... | .................... | .................... | .................... | .................... | .................... |
| Y | 1305 hCoV-19/Switzerland/BE-ETHZ-360337/2020\|EPI_ISL_72 | .................... | .................... | .................... | .................... | ......K............. | .................... |
| Y | 1307 hCoV-19/Switzerland/ZH-ETHZ-360328/2020\|EPI_ISL_72 | .................... | .................... | .................... | .................... | .................... | .................... |
| Y | 1308 hCoV-19/Switzerland/VD-ETHZ-360318/2020\|EPI_ISL_72 | .................... | .................... | .................... | .................... | .................... | .................... |
| Y | 1309 hCoV-19/Switzerland/BE-ETHZ-360317/2020\|EPI_ISL_72 | .................... | .................... | .................... | .................... | .................... | .................... |
| Y | 1310 hCoV-19/Switzerland/VD-ETHZ-360315/2020\|EPI_ISL_72 | .................... | .................... | .................... | .................... | .................... | .................... |
| Y | 1311 hCoV-19/Switzerland/VD-ETHZ-360313/2020\|EPI_ISL_72 | .................... | .................... | .................... | .................... | .................... | .................... |
| Y | 1312 hCoV-19/Switzerland/SH-ETHZ-360310/2020\|EPI_ISL_72 | .................... | .A.................. | .................... | .................... | .................... | .................... |
| Y | 1313 hCoV-19/Switzerland/SH-ETHZ-360305/2020\|EPI_ISL_72 | .................... | .A.................. | .................... | .................... | .................... | .................... |
| Y | 1314 hCoV-19/Switzerland/SO-ETHZ-360303/2020\|EPI_ISL_72 | .................... | .................... | .................... | .................... | .................... | ..K................. |
| Y | 1315 hCoV-19/Switzerland/BE-ETHZ-360300/2020\|EPI_ISL_72 | .................... | .................... | .................... | .................... | .................... | .................... |
| Y | 1316 hCoV-19/Switzerland/VD-ETHZ-360297/2020\|EPI_ISL_72 | .................... | .................... | .................... | .................... | .................... | .................... |
| Y | 1317 hCoV-19/Switzerland/VD-ETHZ-360296/2020\|EPI_ISL_72 | .................... | .................... | .................... | .................... | .................... | .................... |
| Y | 1318 hCoV-19/Switzerland/VD-ETHZ-360295/2020\|EPI_ISL_72 | .................... | .................... | .................... | .................... | .................... | .................... |
| Y | 1319 hCoV-19/Switzerland/BE-ETHZ-360294/2020\|EPI_ISL_72 | .................... | .................... | .................... | .................... | .................... | .................... |
| Y | 1320 hCoV-19/Switzerland/BL-ETHZ-360281/2020\|EPI_ISL_72 | .................... | .................... | .................... | .................... | .................... | .................... |

| | | | | | | | | |
|---|---|---|---|---|---|---|---|---|
| Y | 1321 | hCoV-19/Switzerland/BL-ETHZ-360280/2020\|EPI_ISL_72 | ···················· | ···················· | ···················· | ···················· | ···················· | ···················· |
| Y | 1322 | hCoV-19/Switzerland/BL-ETHZ-360278/2020\|EPI_ISL_72 | ···················· | ···················· | ···················· | ···················· | ···················· | ···················· |
| Y | 1324 | hCoV-19/Switzerland/BL-ETHZ-360275/2020\|EPI_ISL_72 | ···················· | ···················· | ···················· | ···················· | ······K············· | ···················· |
| Y | 1325 | hCoV-19/Switzerland/BL-ETHZ-360274/2020\|EPI_ISL_72 | ···················· | ···················· | ···················· | ···················· | ······K············· | ···················· |
| Y | 1327 | hCoV-19/Switzerland/BL-ETHZ-360267/2020\|EPI_ISL_72 | ···················· | ···················· | ···················· | ···················· | ······K············· | ···················· |
| Y | 1328 | hCoV-19/Switzerland/BL-ETHZ-360266/2020\|EPI_ISL_72 | ···················· | ···················· | ···················· | ···················· | ······K············· | ···················· |
| Y | 1329 | hCoV-19/Switzerland/BL-ETHZ-360265/2020\|EPI_ISL_72 | ···················· | ···················· | ···················· | ···················· | ······K············· | ···················· |
| Y | 1330 | hCoV-19/Switzerland/ZH-ETHZ-360263/2020\|EPI_ISL_72 | ···················· | ···················· | ···················· | ···················· | ···················· | ···················· |
| Y | 1331 | hCoV-19/Switzerland/BE-ETHZ-360261/2020\|EPI_ISL_72 | ···················· | ···················· | ···················· | ···················· | ···················· | ···················· |
| Y | 1333 | hCoV-19/Switzerland/BE-ETHZ-360258/2020\|EPI_ISL_72 | ···················· | ···················· | ···················· | ···················· | ···················· | ···················· |
| Y | 1334 | hCoV-19/Switzerland/AG-ETHZ-360256/2020\|EPI_ISL_72 | ···················· | ···················· | ···················· | ···················· | ···················· | ···················· |
| Y | 1335 | hCoV-19/Switzerland/GR-ETHZ-360253/2020\|EPI_ISL_72 | ···················· | ···················· | ···················· | ···················· | ···················· | ···················· |
| Y | 1336 | hCoV-19/Switzerland/NE-ETHZ-360250/2020\|EPI_ISL_72 | ···················· | ···················· | ···················· | ···················· | ···················· | ···R················ |
| Y | 1339 | hCoV-19/Switzerland/BL-ETHZ-360228/2020\|EPI_ISL_72 | ···················· | ···················· | ···················· | ···················· | ···················· | ···················· |
| Y | 1340 | hCoV-19/Switzerland/AG-ETHZ-360224/2020\|EPI_ISL_72 | ···················· | ···················· | ···················· | ···················· | ···················· | ···················· |
| Y | 1341 | hCoV-19/Switzerland/SO-ETHZ-360222/2020\|EPI_ISL_72 | ···················· | ···················· | ···················· | ···················· | ···················· | ···················· |
| Y | 1342 | hCoV-19/Switzerland/SO-ETHZ-360217/2020\|EPI_ISL_72 | ···················· | ·A·················· | ···················· | ···················· | ···················· | ···················· |
| Y | 1343 | hCoV-19/Switzerland/SO-ETHZ-360216/2020\|EPI_ISL_72 | ···················· | ···················· | ···················· | ···················· | ···················· | ···················· |
| Y | 1344 | hCoV-19/Switzerland/AG-ETHZ-360214/2020\|EPI_ISL_72 | ···················· | ···················· | ···················· | ···················· | ···················· | ···················· |
| Y | 1345 | hCoV-19/Switzerland/SO-ETHZ-360213/2020\|EPI_ISL_72 | ···················· | ···················· | ···················· | ···················· | ···················· | ···················· |
| Y | 1346 | hCoV-19/Switzerland/SO-ETHZ-360212/2020\|EPI_ISL_72 | ···················· | ·A·················· | ···················· | ···················· | ···················· | ···A················ |
| Y | 1347 | hCoV-19/Switzerland/SO-ETHZ-360210/2020\|EPI_ISL_72 | ···················· | ···················· | ···················· | ···················· | ···················· | ···R················ |
| Y | 1348 | hCoV-19/Switzerland/SO-ETHZ-360209/2020\|EPI_ISL_72 | ···················· | ···················· | ···················· | ···················· | ···················· | ···················· |
| Y | 1349 | hCoV-19/Switzerland/NE-ETHZ-350327/2020\|EPI_ISL_72 | ···················· | ···················· | ···················· | ···················· | ···················· | ···················· |
| Y | 1350 | hCoV-19/Switzerland/LU-ETHZ-350450/2020\|EPI_ISL_72 | ···················· | ···················· | ···················· | ···················· | ···················· | ···················· |
| Y | 1351 | hCoV-19/Switzerland/NE-ETHZ-350441/2020\|EPI_ISL_72 | ···················· | ···················· | ···················· | ···················· | ···················· | ···················· |
| Y | 1352 | hCoV-19/Canada/ON-S1913/2020\|EPI_ISL_729265\|2020-1 | ···················· | ·A·················· | ···················· | ···················· | ···················· | ···A················ |
| Y | 1353 | hCoV-19/Canada/ON-S1923/2020\|EPI_ISL_729266\|2020-1 | ···················· | ·A·················· | ···················· | ···················· | ···················· | ···A················ |
| Y | 1354 | hCoV-19/Canada/ON-S1909/2020\|EPI_ISL_729270\|2020-1 | ···················· | ·A·················· | ···················· | ···················· | ···················· | ···A················ |
| Y | 1355 | hCoV-19/Canada/ON-S1799/2020\|EPI_ISL_729271\|2020-1 | ···················· | ···················· | ···················· | ···················· | ···················· | ···················· |
| Y | 1356 | hCoV-19/Canada/ON-S1800/2020\|EPI_ISL_729272\|2020-1 | ···················· | ·A·················· | ···················· | ···················· | ···················· | ···A················ |
| Y | 1357 | hCoV-19/Canada/ON-S1801/2020\|EPI_ISL_729273\|2020-1 | ···················· | ···················· | ···················· | ···················· | ···················· | ···················· |
| Y | 1358 | hCoV-19/Canada/ON-S1802/2020\|EPI_ISL_729274\|2020-1 | ···················· | ·········A·········· | ···················· | ···················· | ···················· | ···················· |
| Y | 1359 | hCoV-19/Canada/ON-S1803/2020\|EPI_ISL_729275\|2020-1 | ···················· | ·········A·········· | ···················· | ···················· | ···················· | ···················· |
| Y | 1360 | hCoV-19/Canada/ON-S1804/2020\|EPI_ISL_729276\|2020-1 | ···················· | ···················· | ···················· | ···················· | ···················· | ···················· |
| Y | 1361 | hCoV-19/Canada/ON-S1805/2020\|EPI_ISL_729277\|2020-1 | ···················· | ···················· | ···················· | ···················· | ···················· | ···················· |
| Y | 1362 | hCoV-19/Canada/ON-S1806/2020\|EPI_ISL_729278\|2020-1 | ···················· | ···················· | ···················· | ···················· | ···················· | ···················· |
| Y | 1363 | hCoV-19/Canada/ON-S1807/2020\|EPI_ISL_729279\|2020-1 | ···················· | ···················· | ···················· | ···················· | ···················· | ···················· |
| Y | 1364 | hCoV-19/Canada/ON-S1826/2020\|EPI_ISL_729290\|2020-1 | ···················· | ·A·················· | ···················· | ···················· | ···················· | ···A················ |
| Y | 1365 | hCoV-19/Canada/ON-S1827/2020\|EPI_ISL_729291\|2020-1 | ···················· | ·A·················· | ···················· | ···················· | ···················· | ···A················ |
| Y | 1366 | hCoV-19/Mexico/BCN-ALSR-4896/2020\|EPI_ISL_730200\|2 | ···················· | ·A·················· | ···················· | ···················· | ···················· | ···················· |
| Y | 1367 | hCoV-19/Mexico/BCN-ALSR-4898/2020\|EPI_ISL_730201\|2 | ···················· | ·A·················· | ···················· | ···················· | ···················· | ···················· |
| Y | 1368 | hCoV-19/Mexico/BCN-ALSR-4899/2020\|EPI_ISL_730202\|2 | ···················· | ·A·················· | ···················· | ···················· | ···················· | ···················· |
| Y | 1369 | hCoV-19/Mexico/BCN-ALSR-4900/2020\|EPI_ISL_730203\|2 | ···················· | ·A·················· | ···················· | ···················· | ···················· | ···················· |
| Y | 1370 | hCoV-19/Mexico/BCN-ALSR-4902/2020\|EPI_ISL_730204\|2 | ···················· | ·A·················· | ···················· | ···················· | ···················· | ···················· |
| Y | 1371 | hCoV-19/Mexico/BCN-ALSR-4903/2020\|EPI_ISL_730205\|2 | ···················· | ·A·················· | ···················· | ···················· | ···················· | ···················· |
| Y | 1372 | hCoV-19/Mexico/BCN-ALSR-4904/2020\|EPI_ISL_730206\|2 | ···················· | ·A·················· | ···················· | ···················· | ···················· | ···················· |
| Y | 1373 | hCoV-19/Mexico/BCN-ALSR-4905/2020\|EPI_ISL_730207\|2 | ···················· | ·A·················· | ···················· | ···················· | ···················· | ···················· |
| Y | 1374 | hCoV-19/Mexico/BCN-ALSR-4910/2020\|EPI_ISL_730208\|2 | ···················· | ·A·················· | ···················· | ···················· | ···················· | ···················· |
| Y | 1375 | hCoV-19/Mexico/BCN-ALSR-4917/2020\|EPI_ISL_730209\|2 | ···················· | ·A·················· | ···················· | ···················· | ···················· | ···················· |
| Y | 1376 | hCoV-19/Mexico/BCN-ALSR-4918/2020\|EPI_ISL_730210\|2 | ···················· | ·A·················· | ···················· | ···················· | ···················· | ···················· |
| Y | 1377 | hCoV-19/Mexico/BCN-ALSR-4920/2020\|EPI_ISL_730211\|2 | ···················· | ···················· | ···················· | ···················· | ···················· | ···················· |
| Y | 1378 | hCoV-19/Mexico/BCN-ALSR-4922/2020\|EPI_ISL_730212\|2 | ···················· | ·A·················· | ···················· | ···················· | ···················· | ···················· |
| Y | 1379 | hCoV-19/Mexico/BCN-ALSR-4923/2020\|EPI_ISL_730213\|2 | ···················· | ·A·················· | ···················· | ···················· | ···················· | ···················· |
| Y | 1380 | hCoV-19/Mexico/BCN-ALSR-4924/2020\|EPI_ISL_730214\|2 | ···················· | ·A·················· | ···················· | ···················· | ···················· | ···················· |
| Y | 1381 | hCoV-19/Mexico/BCN-ALSR-4926/2020\|EPI_ISL_730215\|2 | ···················· | ·A·················· | ···················· | ···················· | ···················· | ···················· |
| Y | 1382 | hCoV-19/Mexico/BCN-ALSR-4927/2020\|EPI_ISL_730216\|2 | ···················· | ·A·················· | ···················· | ···················· | ···················· | ···················· |
| Y | 1383 | hCoV-19/Mexico/BCN-ALSR-4934/2020\|EPI_ISL_730219\|2 | ···················· | ·A·················· | ···················· | ···················· | ···················· | ···················· |
| Y | 1384 | hCoV-19/Mexico/BCN-ALSR-4935/2020\|EPI_ISL_730220\|2 | ···················· | ····G··············· | ···················· | ···················· | ···················· | ···················· |
| Y | 1385 | hCoV-19/Mexico/BCN-ALSR-4937/2020\|EPI_ISL_730221\|2 | ···················· | ·A·················· | ···················· | ···················· | ···················· | ···················· |
| Y | 1386 | hCoV-19/Mexico/BCN-ALSR-4942/2020\|EPI_ISL_730222\|2 | ···················· | ·A·················· | ···················· | ···················· | ···················· | ···················· |
| Y | 1387 | hCoV-19/Mexico/BCN-ALSR-4943/2020\|EPI_ISL_730223\|2 | ···················· | ·A·················· | ···················· | ···················· | ···················· | ···················· |
| Y | 1388 | hCoV-19/Mexico/BCN-ALSR-4947/2020\|EPI_ISL_730225\|2 | ···················· | ·A·················· | ···················· | ···················· | ···················· | ···················· |
| Y | 1389 | hCoV-19/Mexico/BCN-ALSR-4951/2020\|EPI_ISL_730226\|2 | ···················· | ···················· | ···················· | ···················· | ···················· | ···················· |
| Y | 1390 | hCoV-19/Mexico/BCN-ALSR-4952/2020\|EPI_ISL_730227\|2 | ···················· | ·A·················· | ···················· | ···················· | ···················· | ···················· |
| Y | 1391 | hCoV-19/Mexico/BCN-ALSR-4953/2020\|EPI_ISL_730228\|2 | ···················· | ·A·················· | ···················· | ···················· | ···················· | ···················· |
| Y | 1392 | hCoV-19/Jordan/AM-ALSR-4955/2020\|EPI_ISL_730229\|20 | ···················· | ···················· | ···················· | ···················· | ···················· | ···················· |
| Y | 1393 | hCoV-19/Jordan/AM-ALSR-4956/2020\|EPI_ISL_730230\|20 | ···················· | ···················· | ···················· | ···················· | ···················· | ···················· |
| Y | 1394 | hCoV-19/Jordan/AM-ALSR-4957/2020\|EPI_ISL_730231\|20 | ···················· | ···················· | ···················· | ···················· | ···················· | ···················· |
| Y | 1395 | hCoV-19/Jordan/AM-ALSR-4959/2020\|EPI_ISL_730232\|20 | ···················· | ···················· | ···················· | ···················· | ···················· | ···················· |
| Y | 1396 | hCoV-19/Jordan/AM-ALSR-4960/2020\|EPI_ISL_730233\|20 | ···················· | ···················· | ···················· | ···················· | ···················· | ···················· |
| Y | 1397 | hCoV-19/Jordan/AM-ALSR-4961/2020\|EPI_ISL_730234\|20 | ···················· | ·A·················· | ···················· | ···················· | ···················· | ···A················ |
| Y | 1398 | hCoV-19/Jordan/AM-ALSR-4962/2020\|EPI_ISL_730235\|20 | ···················· | ···················· | ···················· | ···················· | ···················· | ···················· |
| Y | 1399 | hCoV-19/Jordan/AM-ALSR-4963/2020\|EPI_ISL_730236\|20 | ···················· | ···················· | ···················· | ···················· | ···················· | ···················· |
| Y | 1400 | hCoV-19/Jordan/AM-ALSR-4964/2020\|EPI_ISL_730237\|20 | ···················· | ···················· | ···················· | ···················· | ···················· | ···················· |
| Y | 1401 | hCoV-19/Jordan/AM-ALSR-4965/2020\|EPI_ISL_730238\|20 | ···················· | ···················· | ···················· | ···················· | ···················· | ···················· |
| Y | 1402 | hCoV-19/Jordan/AM-ALSR-4966/2020\|EPI_ISL_730239\|20 | ···················· | ···················· | ···················· | ···················· | ···················· | ···················· |
| Y | 1403 | hCoV-19/Jordan/AM-ALSR-4967/2020\|EPI_ISL_730240\|20 | ···················· | ···················· | ···················· | ···················· | ···················· | ···················· |
| Y | 1404 | hCoV-19/Jordan/AM-ALSR-4968/2020\|EPI_ISL_730241\|20 | ···················· | ···················· | ···················· | ···················· | ···················· | ···················· |
| Y | 1405 | hCoV-19/Jordan/AM-ALSR-4969/2020\|EPI_ISL_730242\|20 | ···················· | ···················· | ···················· | ···················· | ···················· | ···················· |

| | | | | | | | |
|---|---|---|---|---|---|---|---|
| Y | 1406 hCoV-19/Jordan/AM-ALSR-4970/2020\|EPI_ISL_730243\|20 | ···················· | ·A·················· | ···················· | ···················· | ···················· | ···A················ |
| Y | 1407 hCoV-19/Jordan/AM-ALSR-4971/2020\|EPI_ISL_730244\|20 | ···················· | ···················· | ···················· | ···················· | ···················· | ···················· |
| Y | 1408 hCoV-19/Jordan/AM-ALSR-4972/2020\|EPI_ISL_730245\|20 | ···················· | ···················· | ···················· | ···················· | ···················· | ···················· |
| Y | 1409 hCoV-19/Jordan/AM-ALSR-4973/2020\|EPI_ISL_730246\|20 | ···················· | ···················· | ···················· | ···················· | ···················· | ···················· |
| Y | 1410 hCoV-19/Jordan/AM-ALSR-4974/2020\|EPI_ISL_730247\|20 | ···················· | ···················· | ···················· | ···················· | ···················· | ···················· |
| Y | 1411 hCoV-19/Jordan/AM-ALSR-4975/2020\|EPI_ISL_730248\|20 | ···················· | ···················· | ···················· | ···················· | ···················· | ···················· |
| Y | 1412 hCoV-19/Jordan/AM-ALSR-4976/2020\|EPI_ISL_730249\|20 | ···················· | ···················· | ···················· | ···················· | ···················· | ···················· |
| Y | 1413 hCoV-19/Jordan/AM-ALSR-4977/2020\|EPI_ISL_730250\|20 | ···················· | ···················· | ···················· | ···················· | ······T············· | ···················· |
| Y | 1414 hCoV-19/Jordan/AM-ALSR-4978/2020\|EPI_ISL_730251\|20 | ···················· | ···················· | ···················· | ···················· | ···················· | ···················· |
| Y | 1415 hCoV-19/Jordan/AM-ALSR-4979/2020\|EPI_ISL_730252\|20 | ···················· | ···················· | ···················· | ···················· | ···················· | ···················· |
| Y | 1416 hCoV-19/Jordan/AM-ALSR-4980/2020\|EPI_ISL_730253\|20 | ···················· | ···················· | ···················· | ···················· | ···················· | ···················· |
| Y | 1417 hCoV-19/Jordan/AM-ALSR-4981/2020\|EPI_ISL_730254\|20 | ···················· | ···················· | ···················· | ···················· | ···················· | ···················· |
| Y | 1418 hCoV-19/Jordan/AM-ALSR-4982/2020\|EPI_ISL_730255\|20 | ···················· | ···················· | ···················· | ···················· | ···················· | ···················· |
| Y | 1419 hCoV-19/Jordan/AM-ALSR-4983/2020\|EPI_ISL_730256\|20 | ···················· | ···················· | ···················· | ···················· | ···················· | ···················· |
| Y | 1420 hCoV-19/Jordan/AM-ALSR-4984/2020\|EPI_ISL_730257\|20 | ···················· | ···················· | ···················· | ···················· | ···················· | ···················· |
| Y | 1421 hCoV-19/Jordan/AM-ALSR-4985/2020\|EPI_ISL_730258\|20 | ···················· | ···················· | ···················· | ···················· | ···················· | ···················· |
| Y | 1422 hCoV-19/Jordan/AM-ALSR-4986/2020\|EPI_ISL_730259\|20 | ···················· | ···················· | ···················· | ···················· | ···················· | ···················· |
| Y | 1423 hCoV-19/Jordan/AM-ALSR-4987/2020\|EPI_ISL_730260\|20 | ···················· | ···················· | ···················· | ···················· | ···················· | ···················· |
| Y | 1424 hCoV-19/Jordan/AM-ALSR-4988/2020\|EPI_ISL_730261\|20 | ···················· | ···················· | ···················· | ···················· | ···················· | ···················· |
| Y | 1425 hCoV-19/Jordan/AM-ALSR-4990/2020\|EPI_ISL_730263\|20 | ···················· | ···················· | ···················· | ···················· | ···················· | ···················· |
| Y | 1426 hCoV-19/Jordan/AM-ALSR-4991/2020\|EPI_ISL_730264\|20 | ···················· | ·A·················· | ···················· | ···················· | ···················· | ···A················ |
| Y | 1427 hCoV-19/Jordan/AM-ALSR-4992/2020\|EPI_ISL_730265\|20 | ···················· | ·A·················· | ···················· | ···················· | ···················· | ···A················ |
| Y | 1428 hCoV-19/Jordan/AM-ALSR-4993/2020\|EPI_ISL_730266\|20 | ···················· | ·A·················· | ···················· | ···················· | ···················· | ···A················ |
| Y | 1429 hCoV-19/Jordan/AM-ALSR-4994/2020\|EPI_ISL_730267\|20 | ···················· | ···················· | ···················· | ···················· | ···················· | ···················· |
| Y | 1430 hCoV-19/Jordan/AM-ALSR-4995/2020\|EPI_ISL_730268\|20 | ···················· | ···················· | ···················· | ···················· | ···················· | ···················· |
| Y | 1431 hCoV-19/Jordan/AM-ALSR-4996/2020\|EPI_ISL_730269\|20 | ···················· | ·A·················· | ···················· | ···················· | ···················· | ···A················ |
| Y | 1432 hCoV-19/Jordan/AM-ALSR-5017/2020\|EPI_ISL_730290\|20 | ···················· | ···················· | ···················· | ···················· | ···················· | ···················· |
| Y | 1433 hCoV-19/USA/CA-ALSR-5019/2020\|EPI_ISL_730291\|2020- | ···················· | ·A·················· | ···················· | ···················· | ···················· | ···················· |
| Y | 1434 hCoV-19/USA/CA-ALSR-5025/2020\|EPI_ISL_730296\|2020- | ········T··········· | ·A·················· | ···················· | ···················· | ···················· | ···················· |
| Y | 1435 hCoV-19/USA/CA-ALSR-5026/2020\|EPI_ISL_730297\|2020- | ···················· | ·A·················· | ···················· | ···················· | ···················· | ···················· |
| Y | 1436 hCoV-19/USA/CA-ALSR-5028/2020\|EPI_ISL_730299\|2020- | ···················· | ·A·················· | ···················· | ···················· | ···················· | ···················· |
| Y | 1437 hCoV-19/England/ALDP-C3A696/2020\|EPI_ISL_731514\|20 | ···················· | ···················· | ···················· | ···················· | ···················· | ···················· |
| Y | 1438 hCoV-19/England/ALDP-C397A3/2020\|EPI_ISL_731515\|20 | ···················· | ···················· | ···················· | ···················· | ···················· | ···················· |
| Y | 1439 hCoV-19/England/ALDP-C3AB3D/2020\|EPI_ISL_731516\|20 | ···················· | ···················· | ···················· | ···················· | ···················· | ···················· |
| Y | 1440 hCoV-19/England/QEUH-C34C5E/2020\|EPI_ISL_731517\|20 | ···················· | ···················· | ···················· | ···················· | ···················· | ···················· |
| Y | 1441 hCoV-19/England/ALDP-C394E8/2020\|EPI_ISL_731520\|20 | ···················· | ···················· | ···················· | ···················· | ···················· | ···················· |
| Y | 1442 hCoV-19/England/ALDP-C3AD19/2020\|EPI_ISL_731524\|20 | ···················· | ···················· | ···················· | ···················· | ···················· | ···················· |
| Y | 1443 hCoV-19/England/ALDP-C3A450/2020\|EPI_ISL_731525\|20 | ···················· | ···················· | ···················· | ···················· | ···················· | ···················· |
| Y | 1444 hCoV-19/England/ALDP-C3965B/2020\|EPI_ISL_731526\|20 | ···················· | ···················· | ···················· | ···················· | ······T············· | ···················· |
| Y | 1445 hCoV-19/England/ALDP-C3A1C2/2020\|EPI_ISL_731527\|20 | ···················· | ·A·················· | ···················· | ···················· | ···················· | ···A················ |
| Y | 1446 hCoV-19/England/ALDP-C3AD28/2020\|EPI_ISL_731528\|20 | ···················· | ···················· | ···················· | ···················· | ···················· | ···················· |
| Y | 1447 hCoV-19/England/ALDP-C3A72A/2020\|EPI_ISL_731530\|20 | ···················· | ···················· | ···················· | ···················· | ···················· | ···················· |
| Y | 1448 hCoV-19/England/ALDP-C395B8/2020\|EPI_ISL_731532\|20 | ···················· | ···················· | ···················· | ···················· | ···················· | ···················· |
| Y | 1449 hCoV-19/England/ALDP-C39AF5/2020\|EPI_ISL_731534\|20 | ···················· | ···················· | ···················· | ···················· | ···················· | ···················· |
| Y | 1450 hCoV-19/England/QEUH-C30EC5/2020\|EPI_ISL_731536\|20 | ···················· | ···················· | ···················· | ···················· | ···················· | ···················· |
| Y | 1451 hCoV-19/England/ALDP-C3ABD3/2020\|EPI_ISL_731538\|20 | ···················· | ···················· | ···················· | ···················· | ···················· | ···················· |
| Y | 1452 hCoV-19/England/ALDP-C39354/2020\|EPI_ISL_731539\|20 | ···················· | ···················· | ···················· | ···················· | ···················· | ···················· |
| Y | 1453 hCoV-19/England/ALDP-C3A836/2020\|EPI_ISL_731540\|20 | ···················· | ···················· | ···················· | ···················· | ···················· | ···················· |
| Y | 1454 hCoV-19/England/QEUH-C310A0/2020\|EPI_ISL_731544\|20 | ···················· | ···················· | ···················· | ···················· | ···················· | ···················· |
| Y | 1455 hCoV-19/England/ALDP-C39381/2020\|EPI_ISL_731545\|20 | ···················· | ···················· | ···················· | ···················· | ···················· | ···················· |
| Y | 1456 hCoV-19/England/ALDP-C3AB5B/2020\|EPI_ISL_731547\|20 | ···················· | ·A·················· | ···················· | ···················· | ···················· | ···················· |
| Y | 1457 hCoV-19/England/QEUH-C31671/2020\|EPI_ISL_731550\|20 | ···················· | ···················· | ···················· | ···················· | ···················· | ···················· |
| Y | 1458 hCoV-19/England/QEUH-C31583/2020\|EPI_ISL_731551\|20 | ···················· | ···················· | ···················· | ···················· | ···················· | ···················· |
| Y | 1459 hCoV-19/England/ALDP-C3BB0F/2020\|EPI_ISL_731552\|20 | ···················· | ···················· | ···················· | ···················· | ···················· | ···················· |
| Y | 1460 hCoV-19/England/ALDP-C3BCFD/2020\|EPI_ISL_731553\|20 | ···················· | ···················· | ···················· | ···················· | ···················· | ···················· |
| Y | 1461 hCoV-19/England/ALDP-C3B89F/2020\|EPI_ISL_731554\|20 | ···················· | ···················· | ···················· | ···················· | ···················· | ···················· |
| Y | 1462 hCoV-19/England/ALDP-C3C03C/2020\|EPI_ISL_731558\|20 | ···················· | ···················· | ···················· | ···················· | ···················· | ···················· |
| Y | 1463 hCoV-19/England/ALDP-C3DAB5/2020\|EPI_ISL_731559\|20 | ···················· | ···················· | ···················· | ···················· | ···················· | ···················· |
| Y | 1464 hCoV-19/England/ALDP-C3DA6A/2020\|EPI_ISL_731561\|20 | ···················· | ···················· | ···················· | ···················· | ···················· | ···················· |
| Y | 1465 hCoV-19/England/ALDP-C3D6DF/2020\|EPI_ISL_731562\|20 | ···················· | ···················· | ···················· | ···················· | ···················· | ···················· |
| Y | 1466 hCoV-19/England/ALDP-C3D930/2020\|EPI_ISL_731564\|20 | ···················· | ···················· | ···················· | ···················· | ···················· | ···················· |
| Y | 1467 hCoV-19/England/ALDP-C3BFC7/2020\|EPI_ISL_731566\|20 | ···················· | ···················· | ···················· | ···················· | ···················· | ···················· |
| Y | 1468 hCoV-19/England/ALDP-C3C9C8/2020\|EPI_ISL_731568\|20 | ···················· | ·A·················· | ···················· | ···················· | ···················· | ···A················ |
| Y | 1469 hCoV-19/England/ALDP-C3AAD6/2020\|EPI_ISL_731571\|20 | ···················· | ·A·················· | ···················· | ···················· | ···················· | ···A················ |
| Y | 1470 hCoV-19/England/ALDP-C3C6B2/2020\|EPI_ISL_731572\|20 | ···················· | ···················· | ···················· | ···················· | ···················· | ···················· |
| Y | 1471 hCoV-19/England/ALDP-C3BF7C/2020\|EPI_ISL_731574\|20 | ···················· | ···················· | ···················· | ···················· | ···················· | ···················· |
| Y | 1472 hCoV-19/England/ALDP-C3CB1D/2020\|EPI_ISL_731575\|20 | ···················· | ···················· | ···················· | ···················· | ···················· | ···················· |
| Y | 1473 hCoV-19/England/ALDP-C3B097/2020\|EPI_ISL_731576\|20 | ···················· | ···················· | ···················· | ···················· | ···················· | ···················· |
| Y | 1474 hCoV-19/England/ALDP-C3CE14/2020\|EPI_ISL_731577\|20 | ···················· | ···················· | ···················· | ···················· | ···················· | ···················· |
| Y | 1475 hCoV-19/England/ALDP-C3DD25/2020\|EPI_ISL_731582\|20 | ···················· | ···················· | ···················· | ···················· | ···················· | ···················· |
| Y | 1476 hCoV-19/England/ALDP-C3BFB8/2020\|EPI_ISL_731583\|20 | ···················· | ···················· | ···················· | ···················· | ···················· | ···················· |
| Y | 1477 hCoV-19/England/ALDP-C3DCCE/2020\|EPI_ISL_731587\|20 | ···················· | ···················· | ···················· | ···················· | ···················· | ···················· |
| Y | 1478 hCoV-19/England/ALDP-C3C694/2020\|EPI_ISL_731590\|20 | ···················· | ···················· | ···················· | ···················· | ···················· | ···················· |
| Y | 1479 hCoV-19/England/ALDP-C3B273/2020\|EPI_ISL_731592\|20 | ·····G·············· | ···················· | ···················· | ···················· | ···················· | ···················· |
| Y | 1480 hCoV-19/England/ALDP-C3D45D/2020\|EPI_ISL_731593\|20 | ···················· | ···················· | ···················· | ···················· | ···················· | ···················· |
| Y | 1481 hCoV-19/England/ALDP-C3C88F/2020\|EPI_ISL_731595\|20 | ···················· | ···················· | ···················· | ···················· | ···················· | ···················· |
| Y | 1482 hCoV-19/England/ALDP-C3D6B1/2020\|EPI_ISL_731598\|20 | ···················· | ····A··············· | ···················· | ···················· | ···················· | ···················· |
| Y | 1483 hCoV-19/England/ALDP-C3BEBB/2020\|EPI_ISL_731599\|20 | ···················· | ···················· | ···················· | ···················· | ···················· | ···················· |
| Y | 1484 hCoV-19/Poland/77057/2020\|EPI_ISL_732844\|2020-11 | ···················· | ···················· | ···················· | ···················· | ···················· | ···················· |
| Y | 1485 hCoV-19/Poland/79309/2020\|EPI_ISL_732845\|2020-11 | ···················· | ···················· | ···················· | ···················· | ···················· | ···················· |

Y 1486 hCoV-19/Poland/77075/2020|EPI_ISL_732846|2020-11 ···················· ···················· ···················· ···················· ···················· ····················
Y 1487 hCoV-19/Poland/79284/2020|EPI_ISL_732847|2020-11 ···················· ···················· ···················· ···················· ···················· ····················
Y 1488 hCoV-19/Poland/77178/2020|EPI_ISL_732848|2020-11 ···················· ···················· ···················· ···················· ···················· ····················
Y 1489 hCoV-19/Poland/79329/2020|EPI_ISL_732849|2020-11 ···················· ···················· ···················· ···················· ···················· ····················
Y 1490 hCoV-19/Poland/79302/2020|EPI_ISL_732850|2020-11 ···················· ···················· ···················· ···················· ···················· ····················
Y 1491 hCoV-19/Poland/77245/2020|EPI_ISL_732851|2020-11 ···················· ···················· ···················· ···················· ···················· ····················
Y 1492 hCoV-19/Poland/77142/2020|EPI_ISL_732852|2020-11 ···················· ···················· ···················· ···················· ···················· ····················
Y 1493 hCoV-19/Poland/78588/2020|EPI_ISL_732853|2020-11 ···················· ···················· ···················· ···················· ···················· ····················
Y 1494 hCoV-19/Poland/79299/2020|EPI_ISL_732854|2020-11 ···················· ···················· ···················· ···················· ···················· ····················
Y 1495 hCoV-19/Poland/77741/2020|EPI_ISL_732855|2020-11 ···················· ···················· ···················· ···················· ···················· ····················
Y 1496 hCoV-19/Poland/76078/2020|EPI_ISL_732856|2020-11 ···················· ···················· ···················· ···················· ···················· ····················
Y 1497 hCoV-19/Poland/7629/2020|EPI_ISL_732858|2020-11 ···················· ···················· ···················· ···················· ···················· ····················
Y 1498 hCoV-19/Poland/77061/2020|EPI_ISL_732859|2020-11 ···················· ···················· ···················· ···················· ···················· ····················
Y 1499 hCoV-19/Poland/76089/2020|EPI_ISL_732860|2020-11 ···················· ···················· ···················· ···················· ···················· ····················
Y 1500 hCoV-19/Poland/76331/2020|EPI_ISL_732861|2020-11 ···················· ···················· ···················· ···················· ···················· ····················
Y 1501 hCoV-19/Poland/77242/2020|EPI_ISL_732862|2020-11 ···················· ···················· ···················· ···················· ···················· ····················
Y 1502 hCoV-19/Poland/76330/2020|EPI_ISL_732863|2020-11 ···················· ···················· ···················· ···················· ···················· ····················
Y 1503 hCoV-19/Poland/76235/2020|EPI_ISL_732864|2020-11 ···················· ·A·················· ···················· ···················· ···················· ····················
Y 1504 hCoV-19/Poland/76302/2020|EPI_ISL_732865|2020-11 ···················· ···················· ···················· ···················· ···················· ····················
Y 1505 hCoV-19/Poland/76327/2020|EPI_ISL_732866|2020-11 ···················· ···················· ···················· ···················· ···················· ····················
Y 1506 hCoV-19/Poland/77073/2020|EPI_ISL_732867|2020-11 ···················· ···················· ···················· ···················· ···················· ····················
Y 1507 hCoV-19/Poland/77111/2020|EPI_ISL_732868|2020-11 ···················· ···················· ···················· ···················· ···················· ····················
Y 1508 hCoV-19/Poland/77087/2020|EPI_ISL_732869|2020-11 ···················· ·A·················· ···················· ···················· ···················· ····················
Y 1509 hCoV-19/Poland/77210/2020|EPI_ISL_732870|2020-11 ···················· ···················· ···················· ···················· ···················· ····················
Y 1510 hCoV-19/Poland/76082/2020|EPI_ISL_732871|2020-11 ···················· ···················· ···················· ···················· ···················· ····················
Y 1511 hCoV-19/Poland/76206/2020|EPI_ISL_732872|2020-11 ···················· ·A·················· ···················· ···················· ···················· ····················
Y 1512 hCoV-19/Poland/76023/2020|EPI_ISL_732873|2020-11 ···················· ·A·················· ···················· ···················· ···················· ····················
Y 1513 hCoV-19/Poland/76112/2020|EPI_ISL_732874|2020-11 ···················· ···················· ···················· ···················· ···················· ····················
Y 1514 hCoV-19/Poland/77056/2020|EPI_ISL_732875|2020-11 ···················· ···················· ···················· ···················· ···················· ····················
Y 1515 hCoV-19/Poland/77235/2020|EPI_ISL_732876|2020-11 ···················· ···················· ···················· ···················· ···················· ····················
Y 1516 hCoV-19/Poland/76217/2020|EPI_ISL_732877|2020-11 ···················· ···················· ···················· ···················· ···················· ····················
Y 1517 hCoV-19/Poland/77079/2020|EPI_ISL_732878|2020-11 ···················· ···················· ···················· ···················· ···················· ····················
Y 1518 hCoV-19/Poland/76087/2020|EPI_ISL_732880|2020-11 ···················· ···················· ···················· ···················· ···················· ····················
Y 1519 hCoV-19/Poland/77114/2020|EPI_ISL_732881|2020-11 ···················· ···················· ···················· ···················· ···················· ····················
Y 1520 hCoV-19/Poland/76038/2020|EPI_ISL_732883|2020-11 ···················· ···················· ···················· ···················· ···················· ····················
Y 1521 hCoV-19/Poland/78566/2020|EPI_ISL_732884|2020-11 ···················· ···················· ···················· ···················· ···················· ····················
Y 1522 hCoV-19/Poland/79330/2020|EPI_ISL_732885|2020-11 ···················· ···················· ···················· ···················· ···················· ····················
Y 1523 hCoV-19/Poland/79335/2020|EPI_ISL_732886|2020-11 ···················· ···················· ···················· ···················· ···················· ····················
Y 1524 hCoV-19/Poland/77205/2020|EPI_ISL_732887|2020-11 ···················· ···················· ···················· ···················· ···················· ····················
Y 1525 hCoV-19/Poland/77213/2020|EPI_ISL_732888|2020-11 ···················· ···················· ···················· ···················· ···················· ····················
Y 1526 hCoV-19/Poland/77179/2020|EPI_ISL_732889|2020-11 ···················· ···················· ···················· ···················· ···················· ····················
Y 1527 hCoV-19/Poland/79293/2020|EPI_ISL_732890|2020-11 ···················· ···················· ···················· ···················· ···················· ····················
Y 1528 hCoV-19/Poland/79304/2020|EPI_ISL_732891|2020-11 ···················· ···················· ···················· ···················· ···················· ····················
Y 1529 hCoV-19/Poland/77174/2020|EPI_ISL_732892|2020-11 ···················· ····A··············· ···················· ···················· ···················· ····················
Y 1530 hCoV-19/Poland/79327/2020|EPI_ISL_732893|2020-11 ···················· ···················· ···················· ···················· ···················· ····················
Y 1531 hCoV-19/Poland/79351/2020|EPI_ISL_732894|2020-11 ···················· ···················· ···················· ···················· ···················· ····················
Y 1532 hCoV-19/Poland/79290/2020|EPI_ISL_732895|2020-11 ···················· ···················· ···················· ···················· ···················· ····················
Y 1533 hCoV-19/Poland/79297/2020|EPI_ISL_732896|2020-11 ···················· ···················· ···················· ···················· ···················· ····················
Y 1534 hCoV-19/Poland/77203/2020|EPI_ISL_732897|2020-11 ···················· ···················· ···················· ···················· ···················· ····················
Y 1535 hCoV-19/Poland/78562/2020|EPI_ISL_732898|2020-11 ···················· ···················· ···················· ···················· ···················· ····················
Y 1536 hCoV-19/Poland/78492/2020|EPI_ISL_732899|2020-11 ···················· ···················· ···················· ···················· ···················· ····················
Y 1537 hCoV-19/England/LIVE-DCC12E/2020|EPI_ISL_724490|20 ···················· ···················· ···················· ···················· ···················· ····················
Y 1538 hCoV-19/England/LIVE-DCC258/2020|EPI_ISL_724496|20 ···················· ···················· ···················· ···················· ···A················ ····················
Y 1539 hCoV-19/England/LIVE-DCC30A/2020|EPI_ISL_724499|20 ···················· ···················· ···················· ···················· ···················· ····················
Y 1540 hCoV-19/Wales/PHWC-4928DA/2020|EPI_ISL_725716|2020 ···················· ···················· ···················· ···················· ···················· ····················
Y 1541 hCoV-19/Wales/PHWC-4928E9/2020|EPI_ISL_725717|2020 ···················· ···················· ···················· ···················· ···················· ····················
Y 1542 hCoV-19/Wales/PHWC-492904/2020|EPI_ISL_725719|2020 ···················· ···················· ···················· ···················· ···················· ····················
Y 1543 hCoV-19/Wales/PHWC-492913/2020|EPI_ISL_725720|2020 ···················· ···················· ···················· ···················· ···················· ····················
Y 1544 hCoV-19/Wales/PHWC-492931/2020|EPI_ISL_725722|2020 ···················· ···················· ···················· ···················· ···················· ····················
Y 1546 hCoV-19/Wales/PHWC-49295F/2020|EPI_ISL_725724|2020 ···················· ···················· ···················· ···················· ···················· ····················
Y 1547 hCoV-19/Wales/PHWC-49296E/2020|EPI_ISL_725725|2020 ···················· ···················· ···················· ···················· ···················· ····················
Y 1548 hCoV-19/Wales/PHWC-49297D/2020|EPI_ISL_725726|2020 ···················· ···················· ···················· ···················· ···················· ···R················
Y 1549 hCoV-19/Wales/PHWC-49298C/2020|EPI_ISL_725727|2020 ···················· ···················· ···················· ···················· ···················· ····················
Y 1550 hCoV-19/Wales/PHWC-49299B/2020|EPI_ISL_725728|2020 ···················· ···················· ···················· ···················· ···················· ····················
Y 1551 hCoV-19/Wales/PHWC-4929B9/2020|EPI_ISL_725729|2020 ···················· ···················· ···················· ···················· ···················· ····················
Y 1552 hCoV-19/Wales/PHWC-4929C8/2020|EPI_ISL_725730|2020 ···················· ···················· ···················· ···················· ···················· ····················
Y 1553 hCoV-19/Wales/PHWC-4929E6/2020|EPI_ISL_725732|2020 ···················· ···················· ···················· ···················· ···················· ····················
Y 1554 hCoV-19/Wales/PHWC-4929F5/2020|EPI_ISL_725733|2020 ···················· ···················· ···················· ···················· ···················· ····················
Y 1558 hCoV-19/Wales/PHWC-492A5C/2020|EPI_ISL_725738|2020 ···················· ···················· ···················· ···················· ···················· ····················
Y 1559 hCoV-19/Wales/PHWC-492A6B/2020|EPI_ISL_725739|2020 ···················· ···················· ···················· ···················· ···················· ····················
Y 1560 hCoV-19/Wales/PHWC-492A7A/2020|EPI_ISL_725740|2020 ···················· ···················· ···················· ···················· ···················· ····················
Y 1561 hCoV-19/Wales/PHWC-492A98/2020|EPI_ISL_725742|2020 ···················· ···················· ···················· ···················· ···················· ····················
Y 1562 hCoV-19/Wales/PHWC-492AB6/2020|EPI_ISL_725744|2020 ···················· ···················· ···················· ···················· ···················· ····················
Y 1563 hCoV-19/Wales/PHWC-492AC5/2020|EPI_ISL_725745|2020 ···················· ···················· ···················· ···················· ···················· ····················
Y 1564 hCoV-19/Wales/PHWC-492AD4/2020|EPI_ISL_725746|2020 ···················· ···················· ···················· ···················· ···················· ····················
Y 1565 hCoV-19/Wales/PHWC-492AE3/2020|EPI_ISL_725747|2020 ···················· ···················· ···················· ···················· ···················· ····················
Y 1566 hCoV-19/Wales/PHWC-492AF2/2020|EPI_ISL_725748|2020 ···················· ···················· ···················· ···················· ···················· ····················
Y 1567 hCoV-19/Wales/PHWC-492B1D/2020|EPI_ISL_725750|2020 ···················· ···················· ···················· ···················· ···················· ····················
Y 1568 hCoV-19/Wales/PHWC-492B3B/2020|EPI_ISL_725752|2020 ···················· ······A············· ···················· ···················· ···················· ····················
Y 1569 hCoV-19/Wales/PHWC-492B77/2020|EPI_ISL_725756|2020 ···················· ···················· ···················· ···················· ···················· ····················

| | | | | | | | |
|---|---|---|---|---|---|---|---|
| Y | 1570 hCoV-19/Wales/PHWC-492B86/2020\|EPI_ISL_725757\|2020 | ···················· | ···················· | ····················· | ···················· | ···················· | ···················· |
| Y | 1571 hCoV-19/Wales/PHWC-492BA4/2020\|EPI_ISL_725758\|2020 | ···················· | ···················· | ····················· | ···················· | ···················· | ···················· |
| Y | 1572 hCoV-19/Wales/PHWC-492BC2/2020\|EPI_ISL_725760\|2020 | ···················· | ···················· | ····················· | ···················· | ···················· | ···················· |
| Y | 1573 hCoV-19/Wales/PHWC-492BD1/2020\|EPI_ISL_725761\|2020 | ···················· | ···················· | ····················· | ···················· | ···················· | ···················· |
| Y | 1574 hCoV-19/Wales/PHWC-492BE0/2020\|EPI_ISL_725762\|2020 | ···················· | ···················· | ····················· | ···················· | ···················· | ···················· |
| Y | 1575 hCoV-19/Wales/PHWC-492BFF/2020\|EPI_ISL_725763\|2020 | ···················· | ···················· | ····················· | ···················· | ···················· | ···················· |
| Y | 1576 hCoV-19/Wales/PHWC-492C1A/2020\|EPI_ISL_725765\|2020 | ···················· | ···················· | ····················· | ···················· | ···················· | ···················· |
| Y | 1577 hCoV-19/Wales/PHWC-492C29/2020\|EPI_ISL_725766\|2020 | ···················· | ···················· | ····················· | ···················· | ···················· | ···················· |
| Y | 1578 hCoV-19/Wales/PHWC-492C38/2020\|EPI_ISL_725767\|2020 | ···················· | ···················· | ····················· | ···················· | ···················· | ···················· |
| Y | 1579 hCoV-19/Wales/PHWC-492C56/2020\|EPI_ISL_725768\|2020 | ···················· | ···················· | ····················· | ···················· | ···················· | ···················· |
| Y | 1580 hCoV-19/Wales/PHWC-492C65/2020\|EPI_ISL_725769\|2020 | ···················· | ···················· | ····················· | ···················· | ···················· | ···················· |
| Y | 1581 hCoV-19/Wales/PHWC-492C74/2020\|EPI_ISL_725770\|2020 | ···················· | ···················· | ····················· | ···················· | ···················· | ···················· |
| Y | 1583 hCoV-19/Wales/PHWC-492CA1/2020\|EPI_ISL_725773\|2020 | ···················· | ·····A·············· | ····················· | ···················· | ···················· | ···················· |
| Y | 1584 hCoV-19/Wales/PHWC-492CB0/2020\|EPI_ISL_725774\|2020 | ···················· | ···················· | ····················· | ···················· | ···················· | ···················· |
| Y | 1585 hCoV-19/Wales/PHWC-492CCF/2020\|EPI_ISL_725775\|2020 | ···················· | ···················· | ····················· | ···················· | ···················· | ···················· |
| Y | 1587 hCoV-19/Wales/PHWC-492CED/2020\|EPI_ISL_725777\|2020 | ···················· | ···················· | ····················· | ···················· | ···················· | ···················· |
| Y | 1588 hCoV-19/Wales/PHWC-492CFC/2020\|EPI_ISL_725778\|2020 | ···················· | ···················· | ····················· | ···················· | ···················· | ···················· |
| Y | 1589 hCoV-19/Wales/PHWC-492D26/2020\|EPI_ISL_725780\|2020 | ···················· | ···················· | ····················· | ···················· | ···················· | ···················· |
| Y | 1590 hCoV-19/Wales/PHWC-492D35/2020\|EPI_ISL_725781\|2020 | ···················· | ···················· | ····················· | ···················· | ···················· | ···················· |
| Y | 1593 hCoV-19/Wales/PHWC-492D80/2020\|EPI_ISL_725784\|2020 | ···················· | ···················· | ····················· | ···················· | ···················· | ···················· |
| Y | 1594 hCoV-19/Wales/PHWC-492DAE/2020\|EPI_ISL_725786\|2020 | ···················· | ···················· | ····················· | ···················· | ···················· | ···················· |
| Y | 1595 hCoV-19/Wales/PHWC-492DBD/2020\|EPI_ISL_725787\|2020 | ···················· | ···················· | ····················· | ···················· | ···················· | ···················· |
| Y | 1596 hCoV-19/Wales/PHWC-492DCC/2020\|EPI_ISL_725788\|2020 | ···················· | ···················· | ····················· | ···················· | ···················· | ···················· |
| Y | 1597 hCoV-19/Wales/PHWC-492DDB/2020\|EPI_ISL_725789\|2020 | ···················· | ···················· | ····················· | ···················· | ···················· | ···················· |
| Y | 1598 hCoV-19/Wales/PHWC-492DEA/2020\|EPI_ISL_725790\|2020 | ···················· | ···················· | ····················· | ···················· | ···················· | ···················· |
| Y | 1599 hCoV-19/Wales/PHWC-492DF9/2020\|EPI_ISL_725791\|2020 | ···················· | ···················· | ····················· | ···················· | ···················· | ···················· |
| Y | 1601 hCoV-19/Wales/PHWC-492E14/2020\|EPI_ISL_725793\|2020 | ···················· | ···················· | ····················· | ···················· | ···················· | ···················· |
| Y | 1602 hCoV-19/Wales/PHWC-492E23/2020\|EPI_ISL_725794\|2020 | ···················· | ···················· | ····················· | ···················· | ···················· | ···················· |
| Y | 1603 hCoV-19/Wales/PHWC-492E41/2020\|EPI_ISL_725796\|2020 | ···················· | ·····A·············· | ····················· | ···················· | ···················· | ···················· |
| Y | 1604 hCoV-19/Wales/PHWC-492E6F/2020\|EPI_ISL_725798\|2020 | ···················· | ···················· | ····················· | ···················· | ···················· | ···················· |
| Y | 1605 hCoV-19/Wales/PHWC-492E7E/2020\|EPI_ISL_725799\|2020 | ···················· | ···················· | ····················· | ···················· | ···················· | ···················· |
| Y | 1606 hCoV-19/Canada/ON-S1811/2020\|EPI_ISL_729280\|2020-1 | ···················· | ···················· | ····················· | ···················· | ···················· | ···················· |
| Y | 1607 hCoV-19/Canada/ON-S1812/2020\|EPI_ISL_729281\|2020-1 | ···················· | ···················· | ····················· | ···················· | ···················· | ···················· |
| Y | 1608 hCoV-19/Canada/ON-S1813/2020\|EPI_ISL_729282\|2020-1 | ···················· | ···················· | ····················· | ···················· | ···················· | ···················· |
| Y | 1609 hCoV-19/Canada/ON-S1815/2020\|EPI_ISL_729283\|2020-1 | ···················· | ·A·················· | ····················· | ···················· | ···················· | ···A················ |
| Y | 1610 hCoV-19/Canada/ON-S1820/2020\|EPI_ISL_729284\|2020-1 | ···················· | ·A·················· | ····················· | ···················· | ···················· | ···A················ |
| Y | 1611 hCoV-19/Canada/ON-S1821/2020\|EPI_ISL_729285\|2020-1 | ···················· | ···················· | ····················· | ···················· | ···················· | ···················· |
| Y | 1612 hCoV-19/Canada/ON-S1822/2020\|EPI_ISL_729286\|2020-1 | ···················· | ···················· | ····················· | ···················· | ···················· | ···················· |
| Y | 1613 hCoV-19/Canada/ON-S1823/2020\|EPI_ISL_729287\|2020-1 | ···················· | ···················· | ····················· | ···················· | ···················· | ···················· |
| Y | 1614 hCoV-19/Canada/ON-S1824/2020\|EPI_ISL_729288\|2020-1 | ···················· | ···················· | ····················· | ···················· | ···················· | ···················· |
| Y | 1615 hCoV-19/Canada/ON-S1825/2020\|EPI_ISL_729289\|2020-1 | ···················· | ···················· | ····················· | ···················· | ···················· | ···················· |
| Y | 1616 hCoV-19/Canada/ON-S1828/2020\|EPI_ISL_729292\|2020-1 | ···················· | ·A·················· | ····················· | ···················· | ···················· | ···A················ |
| Y | 1617 hCoV-19/Canada/ON-S1829/2020\|EPI_ISL_729293\|2020-1 | ···················· | ·A·················· | ····················· | ···················· | ···················· | ···A················ |
| Y | 1618 hCoV-19/Canada/ON-S1830/2020\|EPI_ISL_729294\|2020-1 | ···················· | ···················· | ····················· | ···················· | ···················· | ···················· |
| Y | 1619 hCoV-19/Canada/ON-S1831/2020\|EPI_ISL_729295\|2020-1 | ···················· | ···················· | ····················· | ···················· | ···················· | ···················· |
| Y | 1620 hCoV-19/Canada/ON-S1832/2020\|EPI_ISL_729296\|2020-1 | ···················· | ···················· | ····················· | ···················· | ···················· | ···················· |
| Y | 1621 hCoV-19/Canada/ON-S1833/2020\|EPI_ISL_729297\|2020-1 | ···················· | ···················· | ····················· | ···················· | ···················· | ···················· |
| Y | 1622 hCoV-19/Canada/ON-S1834/2020\|EPI_ISL_729298\|2020-1 | ···················· | ·A·················· | ····················· | ···················· | ···················· | ···A················ |
| Y | 1623 hCoV-19/Canada/ON-S1835/2020\|EPI_ISL_729299\|2020-1 | ···················· | ·A·················· | ····················· | ···················· | ···················· | ···A················ |
| Y | 1624 hCoV-19/Jordan/AM-ALSR-4997/2020\|EPI_ISL_730270\|20 | ···················· | ···················· | ····················· | ···················· | ···················· | ···················· |
| Y | 1625 hCoV-19/Jordan/AM-ALSR-4998/2020\|EPI_ISL_730271\|20 | ···················· | ·A·················· | ····················· | ···················· | ···················· | ···A················ |
| Y | 1626 hCoV-19/Jordan/AM-ALSR-4999/2020\|EPI_ISL_730272\|20 | ···················· | ···················· | ····················· | ···················· | ···················· | ···················· |
| Y | 1627 hCoV-19/Jordan/AM-ALSR-5000/2020\|EPI_ISL_730273\|20 | ···················· | ···················· | ····················· | ···················· | ···················· | ···················· |
| Y | 1628 hCoV-19/Jordan/AM-ALSR-5001/2020\|EPI_ISL_730274\|20 | ···················· | ···················· | ····················· | ···················· | ···················· | ···················· |
| Y | 1629 hCoV-19/Jordan/AM-ALSR-5002/2020\|EPI_ISL_730275\|20 | ···················· | ·A·················· | ····················· | ···················· | ···················· | ···A················ |
| Y | 1630 hCoV-19/Jordan/AM-ALSR-5003/2020\|EPI_ISL_730276\|20 | ···················· | ···················· | ····················· | ···················· | ···················· | ···················· |
| Y | 1631 hCoV-19/Jordan/AM-ALSR-5004/2020\|EPI_ISL_730277\|20 | ···················· | ···················· | ····················· | ···················· | ···················· | ···················· |
| Y | 1632 hCoV-19/Jordan/AM-ALSR-5005/2020\|EPI_ISL_730278\|20 | ···················· | ···················· | ····················· | ···················· | ···················· | ···················· |
| Y | 1633 hCoV-19/Jordan/AM-ALSR-5006/2020\|EPI_ISL_730279\|20 | ···················· | ·A·················· | ····················· | ···················· | ···················· | ···A················ |
| Y | 1634 hCoV-19/Jordan/AM-ALSR-5007/2020\|EPI_ISL_730280\|20 | ···················· | ···················· | ····················· | ···················· | ···················· | ···················· |
| Y | 1635 hCoV-19/Jordan/AM-ALSR-5008/2020\|EPI_ISL_730281\|20 | ···················· | ···················· | ····················· | ···················· | ···················· | ···················· |
| Y | 1636 hCoV-19/Jordan/AM-ALSR-5009/2020\|EPI_ISL_730282\|20 | ···················· | ···················· | ····················· | ···················· | ···················· | ···················· |
| Y | 1637 hCoV-19/Jordan/AM-ALSR-5010/2020\|EPI_ISL_730283\|20 | ···················· | ···················· | ····················· | ···················· | ···················· | ···················· |
| Y | 1638 hCoV-19/Jordan/AM-ALSR-5011/2020\|EPI_ISL_730284\|20 | ···················· | ·A·················· | ····················· | ···················· | ···················· | ···A················ |
| Y | 1639 hCoV-19/Jordan/AM-ALSR-5012/2020\|EPI_ISL_730285\|20 | ···················· | ···················· | ····················· | ···················· | ···················· | ···················· |
| Y | 1640 hCoV-19/Jordan/AM-ALSR-5013/2020\|EPI_ISL_730286\|20 | ···················· | ···················· | ····················· | ···················· | ···················· | ···················· |
| Y | 1641 hCoV-19/Jordan/AM-ALSR-5014/2020\|EPI_ISL_730287\|20 | ···················· | ·A·················· | ····················· | ···················· | ···················· | ···A················ |
| Y | 1642 hCoV-19/Jordan/AM-ALSR-5015/2020\|EPI_ISL_730288\|20 | ···················· | ···················· | ····················· | ···················· | ···················· | ···················· |
| Y | 1643 hCoV-19/Jordan/AM-ALSR-5016/2020\|EPI_ISL_730289\|20 | ···················· | ···················· | ····················· | ···················· | ···················· | ···················· |
| Y | 1644 hCoV-19/Belgium/rega-10151120/2020\|EPI_ISL_735100\| | ···················· | ···················· | ····················· | ···················· | ···················· | ···················· |
| Y | 1645 hCoV-19/Belgium/rega-10151121/2020\|EPI_ISL_735101\| | ···················· | ···················· | ····················· | ···················· | ···················· | ···················· |
| Y | 1646 hCoV-19/Belgium/rega-10151183/2020\|EPI_ISL_735102\| | ···················· | ···················· | ····················· | ···················· | ···················· | ···················· |
| Y | 1647 hCoV-19/Belgium/rega-10151184/2020\|EPI_ISL_735103\| | ···················· | ···················· | ····················· | ···················· | ···················· | ···················· |
| Y | 1648 hCoV-19/Belgium/rega-10151185/2020\|EPI_ISL_735104\| | ···················· | ···················· | ····················· | ···················· | ···················· | ···················· |
| Y | 1649 hCoV-19/Belgium/rega-10151186/2020\|EPI_ISL_735105\| | ···················· | ···················· | ····················· | ···················· | ···················· | ···················· |
| Y | 1650 hCoV-19/Belgium/rega-10181066/2020\|EPI_ISL_735106\| | ···················· | ···················· | ····················· | ···················· | ···················· | ···················· |
| Y | 1651 hCoV-19/Belgium/rega-10181067/2020\|EPI_ISL_735107\| | ···················· | ···················· | ····················· | ···················· | ···················· | ···················· |
| Y | 1652 hCoV-19/Belgium/rega-10181068/2020\|EPI_ISL_735108\| | ···················· | ···················· | ····················· | ···················· | ···················· | ···················· |
| Y | 1653 hCoV-19/Belgium/rega-10181069/2020\|EPI_ISL_735109\| | ···················· | ···················· | ····················· | ···················· | ···················· | ···················· |
| Y | 1654 hCoV-19/Belgium/rega-10181070/2020\|EPI_ISL_735110\| | ···················· | ···················· | ····················· | ···················· | ···················· | ···················· |

```
Y 1655 hCoV-19/Belgium/rega-10181071/2020|EPI_ISL_735111| ···················· ···················· ···················· ···················· ···················· ····················
Y 1656 hCoV-19/Belgium/rega-10181072/2020|EPI_ISL_735112| ···················· ···················· ···················· ···················· ···················· ····················
Y 1657 hCoV-19/Belgium/rega-10181073/2020|EPI_ISL_735113| ···················· ···················· ···················· ···················· ···················· ····················
Y 1658 hCoV-19/Belgium/rega-10181074/2020|EPI_ISL_735114| ···················· ···················· ···················· ···················· ···················· ····················
Y 1659 hCoV-19/Belgium/rega-10181075/2020|EPI_ISL_735115| ···················· ···················· ···················· ···················· ···················· ····················
Y 1660 hCoV-19/Belgium/rega-10181076/2020|EPI_ISL_735116| ···················· ···················· ···················· ···················· ···················· ····················
Y 1661 hCoV-19/Belgium/rega-10181077/2020|EPI_ISL_735117| ···················· ···················· ···················· ···················· ···················· ····················
Y 1662 hCoV-19/Belgium/rega-10181078/2020|EPI_ISL_735118| ···················· ···················· ···················· ···················· ···················· ····················
Y 1663 hCoV-19/Belgium/rega-10181079/2020|EPI_ISL_735119| ···················· ···················· ···················· ···················· ···················· ····················
Y 1664 hCoV-19/Belgium/rega-10191103/2020|EPI_ISL_735150| ···················· ···················· ···················· ···················· ···················· ····················
Y 1665 hCoV-19/Belgium/rega-10191107/2020|EPI_ISL_735151| ···················· ···················· ···················· ···················· ···················· ····················
Y 1666 hCoV-19/Belgium/rega-10191109/2020|EPI_ISL_735152| ···················· ···················· ···················· ···················· ···················· ····················
Y 1667 hCoV-19/Belgium/rega-10191122/2020|EPI_ISL_735153| ···················· ···················· ···················· ···················· ······T············· ····················
Y 1668 hCoV-19/Belgium/rega-10191123/2020|EPI_ISL_735154| ···················· ···················· ···················· ···················· ···················· ····················
Y 1669 hCoV-19/Belgium/rega-10191124/2020|EPI_ISL_735155| ···················· ···················· ···················· ···················· ···················· ····················
Y 1670 hCoV-19/Belgium/rega-10191125/2020|EPI_ISL_735156| ···················· ···················· ···················· ···················· ···················· ····················
Y 1671 hCoV-19/Belgium/rega-10191126/2020|EPI_ISL_735157| ···················· ···················· ···················· ···················· ···················· ····················
Y 1672 hCoV-19/Belgium/rega-10191127/2020|EPI_ISL_735158| ···················· ···················· ···················· ···················· ···················· ····················
Y 1673 hCoV-19/Belgium/rega-10191128/2020|EPI_ISL_735159| ···················· ···················· ···················· ···················· ···················· ····················
Y 1674 hCoV-19/England/MILK-C52610/2020|EPI_ISL_736403|20 ···················· ···················· ···················· ···················· ···················· ····················
Y 1675 hCoV-19/England/QEUH-C53F8D/2020|EPI_ISL_736404|20 ···················· ···················· ···················· ···················· ···················· ····················
Y 1676 hCoV-19/England/MILK-C52908/2020|EPI_ISL_736410|20 ···················· ·A·················· ···················· ···················· ···················· ··A·················
Y 1677 hCoV-19/England/QEUH-C54511/2020|EPI_ISL_736414|20 ···················· ···················· ···················· ···················· ···················· ····················
Y 1678 hCoV-19/England/QEUH-C53DFC/2020|EPI_ISL_736416|20 ···················· ···················· ···················· ···················· ···················· ····················
Y 1679 hCoV-19/England/MILK-C52C5A/2020|EPI_ISL_736420|20 ···················· ···················· ···················· ···················· ···················· ····················
Y 1680 hCoV-19/England/MILK-C5264D/2020|EPI_ISL_736421|20 ···················· ···················· ···················· ···················· ···················· ····················
Y 1681 hCoV-19/England/QEUH-C53E71/2020|EPI_ISL_736423|20 ···················· ···················· ···················· ···················· ···················· ····················
Y 1682 hCoV-19/England/QEUH-C5462D/2020|EPI_ISL_736426|20 ···················· ···················· ···················· ···················· ···················· ····················
Y 1683 hCoV-19/England/QEUH-C545E4/2020|EPI_ISL_736431|20 ···················· ···················· ···················· ···················· ···················· ····················
Y 1684 hCoV-19/England/MILK-C52926/2020|EPI_ISL_736433|20 ···················· ·A·················· ···················· ···················· ···················· ··A·················
Y 1685 hCoV-19/England/MILK-C52EFA/2020|EPI_ISL_736435|20 ···················· ···················· ···················· ···················· ···················· ····················
Y 1686 hCoV-19/England/MILK-C52DFD/2020|EPI_ISL_736436|20 ···················· ···················· ···················· ···················· ···················· ····················
Y 1687 hCoV-19/England/QEUH-C54089/2020|EPI_ISL_736437|20 ···················· ···················· ···················· ···················· ···················· ····················
Y 1688 hCoV-19/England/QEUH-C541E0/2020|EPI_ISL_736438|20 ···················· ···················· ···················· ···················· ···················· ····················
Y 1689 hCoV-19/England/QEUH-C5421A/2020|EPI_ISL_736442|20 ···················· ···················· ···················· ···················· ···················· ····················
Y 1690 hCoV-19/England/QEUH-C57B3A/2020|EPI_ISL_736444|20 ···················· ···················· ···················· ···················· ···················· ····················
Y 1691 hCoV-19/England/MILK-C52391/2020|EPI_ISL_736449|20 ···················· ···················· ···················· ···················· ···················· ····················
Y 1692 hCoV-19/England/ALDP-C48D81/2020|EPI_ISL_736482|20 ···················· ···················· ···················· ···················· ···················· ····················
Y 1693 hCoV-19/England/ALDP-C4864A/2020|EPI_ISL_736484|20 ···················· ···················· ···················· ···················· ···················· ····················
Y 1694 hCoV-19/England/ALDP-C484D7/2020|EPI_ISL_736485|20 ···················· ···················· ···················· ···················· ···················· ····················
Y 1695 hCoV-19/England/QEUH-C4BB1B/2020|EPI_ISL_736488|20 ···················· ···················· ···················· ···················· ···················· ····················
Y 1697 hCoV-19/Switzerland/BE-ETHZ-321088/2020|EPI_ISL_73 ···················· ···················· ···················· ···················· ···················· ····················
Y 1698 hCoV-19/Switzerland/BE-ETHZ-321089/2020|EPI_ISL_73 ···················· ···················· ···················· ···················· ···················· ····················
Y 1699 hCoV-19/Switzerland/BE-ETHZ-321090/2020|EPI_ISL_73 ···················· ···················· ···················· ···················· ···················· ····················
Y 1700 hCoV-19/Switzerland/BE-ETHZ-321091/2020|EPI_ISL_73 ···················· ···················· ···················· ···················· ···················· ····················
Y 1701 hCoV-19/Switzerland/BE-ETHZ-321092/2020|EPI_ISL_73 ···················· ···················· ···················· ···················· ···················· ····················
Y 1702 hCoV-19/Switzerland/BE-ETHZ-321094/2020|EPI_ISL_73 ···················· ···················· ···················· ···················· ···················· ····················
Y 1703 hCoV-19/Switzerland/BE-ETHZ-360096/2020|EPI_ISL_73 ···················· ···················· ···················· ···················· ···················· ····················
Y 1704 hCoV-19/Switzerland/BE-ETHZ-410066/2020|EPI_ISL_73 ···················· ···················· ···················· ···················· ···················· ····················
Y 1705 hCoV-19/Switzerland/BE-ETHZ-410142/2020|EPI_ISL_73 ···················· ···················· ···················· ···················· ···················· ····················
Y 1706 hCoV-19/Switzerland/BE-ETHZ-410178/2020|EPI_ISL_73 ···················· ···················· ···················· ···················· ···················· ····················
Y 1707 hCoV-19/Switzerland/BE-ETHZ-410183/2020|EPI_ISL_73 ···················· ···················· ···················· ···················· ···················· ····················
Y 1708 hCoV-19/Switzerland/BE-ETHZ-410185/2020|EPI_ISL_73 ···················· ···················· ···················· ···················· ···················· ····················
Y 1709 hCoV-19/Switzerland/BE-ETHZ-410198/2020|EPI_ISL_73 ···················· ···················· ···················· ···················· ···················· ····················
Y 1711 hCoV-19/Switzerland/BE-ETHZ-410200/2020|EPI_ISL_73 ···················· ···················· ···················· ···················· ···················· ··················Y· 1
Y 1712 hCoV-19/Switzerland/BE-ETHZ-410201/2020|EPI_ISL_73 ···················· ···················· ···················· ···················· ···················· ····················
Y 1714 hCoV-19/Switzerland/BE-ETHZ-410205/2020|EPI_ISL_73 ···················· ···················· ···················· ···················· ···················· ····················
Y 1715 hCoV-19/Switzerland/BE-ETHZ-410206/2020|EPI_ISL_73 ···················· ···················· ···················· ···················· ···················· ····················
Y 1716 hCoV-19/Switzerland/BE-ETHZ-410207/2020|EPI_ISL_73 ···················· ···················· ···················· ···················· ···················· ····················
Y 1717 hCoV-19/Switzerland/BE-ETHZ-410209/2020|EPI_ISL_73 ···················· ···················· ···················· ···················· ···················· ····················
Y 1718 hCoV-19/Switzerland/BE-ETHZ-410210/2020|EPI_ISL_73 ···················· ···················· ···················· ···················· ···················· ····················
Y 1719 hCoV-19/Switzerland/BE-ETHZ-410211/2020|EPI_ISL_73 ···················· ···················· ···················· ···················· ·······S············ ····················
Y 1720 hCoV-19/Switzerland/BE-ETHZ-410212/2020|EPI_ISL_73 ···················· ···················· ···················· ···················· ···················· ····················
Y 1721 hCoV-19/Switzerland/BE-ETHZ-410220/2020|EPI_ISL_73 ···················· ···················· ···················· ···················· ···················· ····················
Y 1722 hCoV-19/Switzerland/BE-ETHZ-410233/2020|EPI_ISL_73 ···················· ···················· ···················· ···················· ···················· ····················
Y 1723 hCoV-19/Switzerland/BE-ETHZ-410235/2020|EPI_ISL_73 ···················· ···················· ···················· ···················· ···················· ····················
Y 1724 hCoV-19/Switzerland/BE-ETHZ-410240/2020|EPI_ISL_73 ···················· ···················· ···················· ···················· ···················· ····················
Y 1725 hCoV-19/Switzerland/BE-ETHZ-410242/2020|EPI_ISL_73 ···················· ···················· ···················· ···················· ···················· ····················
Y 1726 hCoV-19/Switzerland/BE-ETHZ-410250/2020|EPI_ISL_73 ···················· ···················· ···················· ···················· ···················· ····················
Y 1728 hCoV-19/Switzerland/BE-ETHZ-410556/2020|EPI_ISL_73 ···················· ···················· ···················· ···················· ···················· ····················
Y 1729 hCoV-19/Switzerland/BE-ETHZ-410558/2020|EPI_ISL_73 ···················· ···················· ···················· ···················· ···················· ····················
Y 1730 hCoV-19/Switzerland/BE-ETHZ-410561/2020|EPI_ISL_73 ···················· ···················· ···················· ···················· ···················· ····················
Y 1732 hCoV-19/Switzerland/BE-ETHZ-410564/2020|EPI_ISL_73 ···················· ···················· ···················· ···················· ···················· ····················
Y 1733 hCoV-19/Switzerland/BE-ETHZ-410566/2020|EPI_ISL_73 ···················· ···················· ···················· ···················· ···················· ····················
Y 1734 hCoV-19/Switzerland/BE-ETHZ-410569/2020|EPI_ISL_73 ···················· ···················· ···················· ···················· ···················· ····················
Y 1735 hCoV-19/Switzerland/BE-ETHZ-410739/2020|EPI_ISL_73 ···················· ···················· ···················· ···················· ···················· ····················
Y 1736 hCoV-19/Switzerland/BE-ETHZ-410794/2020|EPI_ISL_73 ···················· ···················· ···················· ···················· ···················· ····················
Y 1737 hCoV-19/Switzerland/BE-ETHZ-410875/2020|EPI_ISL_73 ···················· ···················· ···················· ···················· ···················· ····················
Y 1738 hCoV-19/Switzerland/BE-ETHZ-410877/2020|EPI_ISL_73 ···················· ···················· ···················· ···················· ···················· ····················
Y 1739 hCoV-19/Switzerland/BE-ETHZ-410879/2020|EPI_ISL_73 ···················· ···················· ···················· ···················· ···················· ····················
```

| | | | | | | | |
|---|---|---|---|---|---|---|---|
| Y | 1740 hCoV-19/Switzerland/BE-ETHZ-410880/2020\|EPI_ISL_73 | ···················· | ···················· | ···················· | ···················· | ···················· | ···················· |
| Y | 1741 hCoV-19/Switzerland/BE-ETHZ-410882/2020\|EPI_ISL_73 | ···················· | ···················· | ···················· | ···················· | ······K············· | ···················· |
| Y | 1742 hCoV-19/Switzerland/BE-ETHZ-410889/2020\|EPI_ISL_73 | ···················· | ···················· | ···················· | ···················· | ···················· | ···················· |
| Y | 1743 hCoV-19/Switzerland/BE-ETHZ-410902/2020\|EPI_ISL_73 | ···················· | ···················· | ···················· | ···················· | ···················· | ···················· |
| Y | 1744 hCoV-19/Switzerland/BE-ETHZ-410918/2020\|EPI_ISL_73 | ···················· | ···················· | ···················· | ···················· | ···················· | ···················· |
| Y | 1745 hCoV-19/Switzerland/BE-ETHZ-410919/2020\|EPI_ISL_73 | ···················· | ···················· | ···················· | ···················· | ···················· | ···················· |
| Y | 1746 hCoV-19/Switzerland/BE-ETHZ-410920/2020\|EPI_ISL_73 | ···················· | ···················· | ···················· | ···················· | ···················· | ···················· |
| Y | 1747 hCoV-19/Switzerland/GE-ETHZ-320577/2020\|EPI_ISL_73 | ···················· | ···················· | ···················· | ···················· | ···················· | ···················· |
| Y | 1748 hCoV-19/Switzerland/GE-ETHZ-320584/2020\|EPI_ISL_73 | ···················· | ···················· | ···················· | ···················· | ···················· | ···················· |
| Y | 1749 hCoV-19/Switzerland/GE-ETHZ-320595/2020\|EPI_ISL_73 | ···················· | ···················· | ···················· | ···················· | ···················· | ···················· |
| Y | 1750 hCoV-19/Switzerland/GE-ETHZ-320596/2020\|EPI_ISL_73 | ···················· | ···················· | ···················· | ···················· | ···················· | ···················· |
| Y | 1751 hCoV-19/Switzerland/GE-ETHZ-320597/2020\|EPI_ISL_73 | ···················· | ···················· | ···················· | ···················· | ······K············· | ···················· |
| Y | 1752 hCoV-19/Switzerland/GE-ETHZ-320598/2020\|EPI_ISL_73 | ···················· | ···················· | ···················· | ···················· | ···················· | ···················· |
| Y | 1753 hCoV-19/Switzerland/GE-ETHZ-320601/2020\|EPI_ISL_73 | ···················· | ···················· | ···················· | ···················· | ······T············· | ···················· |
| Y | 1754 hCoV-19/Switzerland/GE-ETHZ-410908/2020\|EPI_ISL_73 | ···················· | ···················· | ···················· | ···················· | ···················· | ···················· |
| Y | 1755 hCoV-19/Switzerland/GR-ETHZ-410167/2020\|EPI_ISL_73 | ···················· | ···················· | ···················· | ···················· | ···················· | ···················· |
| Y | 1756 hCoV-19/Switzerland/GR-ETHZ-410168/2020\|EPI_ISL_73 | ···················· | ···················· | ···················· | ···················· | ···················· | ···················· |
| Y | 1757 hCoV-19/Switzerland/GR-ETHZ-410247/2020\|EPI_ISL_73 | ···················· | ···················· | ···················· | ···················· | ···················· | ···················· |
| Y | 1758 hCoV-19/Switzerland/GR-ETHZ-410909/2020\|EPI_ISL_73 | ···················· | ···················· | ···················· | ···················· | ···················· | ···················· |
| Y | 1759 hCoV-19/Switzerland/LU-ETHZ-320593/2020\|EPI_ISL_73 | ···················· | ···················· | ···················· | ···················· | ···················· | ···················· |
| Y | 1760 hCoV-19/Switzerland/LU-ETHZ-320594/2020\|EPI_ISL_73 | ···················· | ···················· | ···················· | ···················· | ···················· | ···················· |
| Y | 1761 hCoV-19/Switzerland/LU-ETHZ-410187/2020\|EPI_ISL_73 | ···················· | ···················· | ···················· | ···················· | ···················· | ···················· |
| Y | 1762 hCoV-19/Switzerland/LU-ETHZ-410188/2020\|EPI_ISL_73 | ···················· | ···················· | ···················· | ···················· | ···················· | ···················· |
| Y | 1763 hCoV-19/Switzerland/NE-ETHZ-410230/2020\|EPI_ISL_73 | ···················· | ···················· | ···················· | ···················· | ···················· | ···················· |
| Y | 1764 hCoV-19/Switzerland/NE-ETHZ-410231/2020\|EPI_ISL_73 | ···················· | ···················· | ···················· | ···················· | ···················· | ···················· |
| Y | 1765 hCoV-19/Switzerland/OW-ETHZ-410147/2020\|EPI_ISL_73 | ···················· | ···················· | ···················· | ···················· | ···················· | ···················· |
| Y | 1766 hCoV-19/Switzerland/SH-ETHZ-410001/2020\|EPI_ISL_73 | ···················· | ···················· | ···················· | ···················· | ···················· | ···················· |
| Y | 1767 hCoV-19/Switzerland/SH-ETHZ-410085/2020\|EPI_ISL_73 | ···················· | ···················· | ···················· | ···················· | ···················· | ···················· |
| Y | 1768 hCoV-19/Switzerland/SH-ETHZ-410241/2020\|EPI_ISL_73 | ···················· | ···················· | ···················· | ···················· | ···················· | ···················· |
| Y | 1769 hCoV-19/Switzerland/SH-ETHZ-410531/2020\|EPI_ISL_73 | ···················· | ·A·················· | ···················· | ···················· | ···················· | ···················· |
| Y | 1770 hCoV-19/Switzerland/SH-ETHZ-410532/2020\|EPI_ISL_73 | ···················· | ···················· | ···················· | ···················· | ···················· | ···················· |
| Y | 1771 hCoV-19/Switzerland/SH-ETHZ-410533/2020\|EPI_ISL_73 | ···················· | ···················· | ···················· | ···················· | ···················· | ···················· |
| Y | 1772 hCoV-19/Switzerland/SH-ETHZ-410535/2020\|EPI_ISL_73 | ···················· | ···················· | ···················· | ···················· | ···················· | ···················· |
| Y | 1773 hCoV-19/Switzerland/SH-ETHZ-410536/2020\|EPI_ISL_73 | ···················· | ···················· | ···················· | ···················· | ···················· | R··················· |
| Y | 1774 hCoV-19/Switzerland/SH-ETHZ-410538/2020\|EPI_ISL_73 | ···················· | ···················· | ···················· | ···················· | ···················· | ···················· |
| Y | 1776 hCoV-19/USA/NC-CDC-2-3693770/2020\|EPI_ISL_747120\|2 | ···················· | ···················· | ···················· | ···················· | ···················· | ···················· |
| Y | 1777 hCoV-19/USA/AZ-CDC-2-3693700/2020\|EPI_ISL_747122\|2 | ···················· | ···················· | ···················· | ···················· | ···················· | ···················· |
| Y | 1778 hCoV-19/USA/AZ-CDC-2-3693477/2020\|EPI_ISL_747123\|2 | ···················· | ·············A······ | ···················· | ···················· | ···················· | ···················· |
| Y | 1779 hCoV-19/USA/AZ-CDC-2-3693553/2020\|EPI_ISL_747124\|2 | ···················· | ···················· | ···················· | ···················· | ···················· | ···················· |
| Y | 1780 hCoV-19/USA/AZ-CDC-2-3693500/2020\|EPI_ISL_747125\|2 | ···················· | ···················· | ···················· | ···················· | ···················· | ···················· |
| Y | 1781 hCoV-19/USA/AZ-CDC-2-3693598/2020\|EPI_ISL_747126\|2 | ···················· | ···················· | ···················· | ···················· | ···················· | ···················· |
| Y | 1782 hCoV-19/USA/OH-CDC-2-3693484/2020\|EPI_ISL_747128\|2 | ···················· | ···················· | ···················· | ···················· | ···················· | ···················· |
| Y | 1783 hCoV-19/USA/OH-CDC-2-3693576/2020\|EPI_ISL_747129\|2 | ···················· | ···················· | ···················· | ···················· | ···················· | ···················· |
| Y | 1784 hCoV-19/USA/OH-CDC-2-3693476/2020\|EPI_ISL_747131\|2 | ···················· | ···················· | ···················· | ···················· | ···················· | ···················· |
| Y | 1785 hCoV-19/USA/CT-CDC-2-3693584/2020\|EPI_ISL_747132\|2 | ···················· | ···················· | ···················· | ···················· | ···················· | ···················· |
| Y | 1786 hCoV-19/USA/CT-CDC-2-3693575/2020\|EPI_ISL_747133\|2 | ···················· | ···················· | ···················· | ···················· | ···················· | ···················· |
| Y | 1787 hCoV-19/USA/CT-CDC-2-3693591/2020\|EPI_ISL_747134\|2 | ···················· | ···················· | ···················· | ···················· | ···················· | ···················· |
| Y | 1788 hCoV-19/USA/CT-CDC-2-3693464/2020\|EPI_ISL_747135\|2 | ···················· | ···················· | ···················· | ···················· | ···················· | ···················· |
| Y | 1789 hCoV-19/USA/KS-CDC-2-3693494/2020\|EPI_ISL_747136\|2 | ···················· | ···················· | ···················· | ···················· | ···················· | ···················· |
| Y | 1790 hCoV-19/USA/KS-CDC-2-3693574/2020\|EPI_ISL_747137\|2 | ···················· | ···················· | ···················· | ···················· | ···················· | ···················· |
| Y | 1791 hCoV-19/USA/KS-CDC-2-3693489/2020\|EPI_ISL_747138\|2 | ···················· | ···················· | ···················· | ···················· | ···················· | ···················· |
| Y | 1792 hCoV-19/USA/KS-CDC-2-3693594/2020\|EPI_ISL_747139\|2 | ···················· | ···················· | ···················· | ···················· | ···················· | ···················· |
| Y | 1793 hCoV-19/England/LIVE-DCC11F/2020\|EPI_ISL_724489\|20 | ···················· | ···················· | ···················· | ···················· | ···················· | ···················· |
| Y | 1795 hCoV-19/USA/WI-UW-2250/2020\|EPI_ISL_728019\|2020-09 | ···················· | ···················· | ···················· | ···················· | ···················· | ···················· |
| Y | 1796 hCoV-19/Canada/ON-S1836/2020\|EPI_ISL_729300\|2020-1 | ···················· | ···················· | ···················· | ···················· | ···················· | ···················· |
| Y | 1797 hCoV-19/Canada/ON-S1837/2020\|EPI_ISL_729301\|2020-1 | ···················· | ·A·················· | ···················· | ···················· | ···················· | ··A················· |
| Y | 1798 hCoV-19/Canada/ON-S1838/2020\|EPI_ISL_729302\|2020-1 | ···················· | ·A·················· | ···················· | ···················· | ···················· | ··A················· |
| Y | 1799 hCoV-19/Canada/ON-S1840/2020\|EPI_ISL_729303\|2020-1 | ···················· | ···················· | ···················· | ···················· | ···················· | ···················· |
| Y | 1800 hCoV-19/Canada/ON-S1842/2020\|EPI_ISL_729304\|2020-1 | ···················· | ·A·················· | ···················· | ···················· | ···················· | ··A················· |
| Y | 1801 hCoV-19/Canada/ON-S1844/2020\|EPI_ISL_729305\|2020-1 | ···················· | ·A·················· | ···················· | ···················· | ···················· | ··A················· |
| Y | 1802 hCoV-19/Canada/ON-S1846/2020\|EPI_ISL_729306\|2020-1 | ···················· | ·A·················· | ···················· | ···················· | ···················· | ··A················· |
| Y | 1803 hCoV-19/Canada/ON-S1847/2020\|EPI_ISL_729307\|2020-1 | ···················· | ···················· | ···················· | ···················· | ···················· | ···················· |
| Y | 1804 hCoV-19/Canada/ON-S1850/2020\|EPI_ISL_729308\|2020-1 | ···················· | ···················· | ···················· | ···················· | ···················· | ···················· |
| Y | 1805 hCoV-19/Canada/ON-S1851/2020\|EPI_ISL_729309\|2020-1 | ···················· | ···················· | ···················· | ···················· | ···················· | ···················· |
| Y | 1806 hCoV-19/Canada/ON-S1852/2020\|EPI_ISL_729310\|2020-1 | ···················· | ···················· | ···················· | ···················· | ···················· | ···················· |
| Y | 1807 hCoV-19/Canada/ON-S1886/2020\|EPI_ISL_729311\|2020-1 | ···················· | ···················· | ···················· | ···················· | ···················· | ···················· |
| Y | 1808 hCoV-19/Canada/ON-S1887/2020\|EPI_ISL_729312\|2020-1 | ···················· | ···················· | ···················· | ···················· | ···················· | ···················· |
| Y | 1809 hCoV-19/Canada/ON-S1888/2020\|EPI_ISL_729313\|2020-1 | ···················· | ···················· | ···················· | ···················· | ···················· | ···················· |
| Y | 1810 hCoV-19/Canada/ON-S1890/2020\|EPI_ISL_729314\|2020-1 | ···················· | ·A·················· | ···················· | ···················· | ···················· | ··A················· |
| Y | 1811 hCoV-19/Canada/ON-S1892/2020\|EPI_ISL_729315\|2020-1 | ···················· | ···················· | ···················· | ···················· | ···················· | ···················· |
| Y | 1812 hCoV-19/Canada/ON-S1893/2020\|EPI_ISL_729316\|2020-1 | ···················· | ·A·················· | ···················· | ···················· | ···················· | ··A················· |
| Y | 1813 hCoV-19/Canada/ON-S1900/2020\|EPI_ISL_729317\|2020-1 | ···················· | ···················· | ···················· | ···················· | ···················· | ···················· |
| Y | 1814 hCoV-19/Canada/ON-S1901/2020\|EPI_ISL_729318\|2020-1 | ···················· | ·A·················· | ···················· | ···················· | ···················· | ··A················· |
| Y | 1815 hCoV-19/Canada/ON-S1903/2020\|EPI_ISL_729319\|2020-1 | ···················· | ···················· | ···················· | ···················· | ···················· | ···················· |
| Y | 1816 hCoV-19/USA/CA-ALSR-5029/2020\|EPI_ISL_730300\|2020- | ···················· | ·A·················· | ···················· | ···················· | ···················· | ···················· |
| Y | 1817 hCoV-19/England/ALDP-C3D53C/2020\|EPI_ISL_731600\|20 | ···················· | ···················· | ···················· | ···················· | ···················· | ···················· |
| Y | 1818 hCoV-19/England/ALDP-C3D596/2020\|EPI_ISL_731601\|20 | ···················· | ···················· | ···················· | ···················· | ···················· | ···················· |
| Y | 1819 hCoV-19/England/ALDP-C3D2F9/2020\|EPI_ISL_731602\|20 | ···················· | ···················· | ···················· | ···················· | ···················· | ···················· |
| Y | 1820 hCoV-19/England/ALDP-C3DCBF/2020\|EPI_ISL_731604\|20 | ···················· | ···················· | ···················· | ···················· | ···················· | ···················· |
| Y | 1821 hCoV-19/England/ALDP-C3BB2D/2020\|EPI_ISL_731605\|20 | ···················· | ···················· | ···················· | ···················· | ···················· | ···················· |

| | | | | | | | |
|---|---|---|---|---|---|---|---|
| Y | 1822 hCoV-19/England/ALDP-C3D37E/2020\|EPI_ISL_731606\|20 | ···················· | ···················· | ···················· | ···················· | ···················· | ···················· |
| Y | 1823 hCoV-19/England/ALDP-C3CA89/2020\|EPI_ISL_731607\|20 | ···················· | ···················· | ···················· | ···················· | ···················· | ···················· |
| Y | 1824 hCoV-19/England/ALDP-C3C70A/2020\|EPI_ISL_731608\|20 | ···················· | ···················· | ···················· | ···················· | ···················· | ···················· |
| Y | 1825 hCoV-19/England/ALDP-C3D350/2020\|EPI_ISL_731610\|20 | ···················· | ···················· | ···················· | ···················· | ···················· | ···················· |
| Y | 1826 hCoV-19/England/ALDP-C3B176/2020\|EPI_ISL_731611\|20 | ···················· | ···················· | ···················· | ···················· | ······T············· | ···················· |
| Y | 1827 hCoV-19/England/ALDP-C3D52D/2020\|EPI_ISL_731613\|20 | ···················· | ···················· | ···················· | ···················· | ···················· | ···················· |
| Y | 1828 hCoV-19/England/ALDP-C3DE13/2020\|EPI_ISL_731614\|20 | ···················· | ···················· | ···················· | ···················· | ···················· | ···················· |
| Y | 1829 hCoV-19/England/ALDP-C3CE6F/2020\|EPI_ISL_731618\|20 | ···················· | ···················· | ···················· | ···················· | ···················· | ···················· |
| Y | 1830 hCoV-19/England/ALDP-C3D0E0/2020\|EPI_ISL_731619\|20 | ···················· | ···················· | ···················· | ···················· | ···················· | ···················· |
| Y | 1831 hCoV-19/England/ALDP-C3C0A5/2020\|EPI_ISL_731622\|20 | ···················· | ·A·················· | ···················· | ···················· | ···················· | ··A················· |
| Y | 1832 hCoV-19/England/ALDP-C3CDBD/2020\|EPI_ISL_731623\|20 | ···················· | ···················· | ···················· | ···················· | ···················· | ···················· |
| Y | 1833 hCoV-19/England/ALDP-C3CDDB/2020\|EPI_ISL_731625\|20 | ···················· | ·A·················· | ···················· | ···················· | ···················· | ··A················· |
| Y | 1834 hCoV-19/England/ALDP-C3AC49/2020\|EPI_ISL_731626\|20 | ···················· | ···················· | ···················· | ···················· | ···················· | ···················· |
| Y | 1835 hCoV-19/England/ALDP-C3DCFB/2020\|EPI_ISL_731630\|20 | ···················· | ···················· | ···················· | ···················· | ···················· | ···················· |
| Y | 1836 hCoV-19/England/ALDP-C3BF12/2020\|EPI_ISL_731631\|20 | ···················· | ···················· | ···················· | ···················· | ···················· | ···················· |
| Y | 1837 hCoV-19/England/ALDP-C3AA7C/2020\|EPI_ISL_731632\|20 | ···················· | ·A·················· | ···················· | ···················· | ···················· | ··A················· |
| Y | 1838 hCoV-19/England/ALDP-C3BAE4/2020\|EPI_ISL_731633\|20 | ···················· | ···················· | ···················· | ···················· | ···················· | ···················· |
| Y | 1839 hCoV-19/Poland/77209/2020\|EPI_ISL_732900\|2020-11 | ···················· | ·A·················· | ···················· | ···················· | ···················· | ···················· |
| Y | 1840 hCoV-19/Poland/78428/2020\|EPI_ISL_732901\|2020-11 | ···················· | ···················· | ···················· | ···················· | ···················· | ···················· |
| Y | 1841 hCoV-19/Poland/77048/2020\|EPI_ISL_732902\|2020-11 | ···················· | ···················· | ···················· | ···················· | ···················· | ···················· |
| Y | 1842 hCoV-19/Poland/77140/2020\|EPI_ISL_732903\|2020-11 | ···················· | ···················· | ···················· | ···················· | ···················· | ···················· |
| Y | 1843 hCoV-19/Poland/78565/2020\|EPI_ISL_732904\|2020-11 | ···················· | ···················· | ···················· | ···················· | ···················· | ···················· |
| Y | 1844 hCoV-19/Poland/79267/2020\|EPI_ISL_732905\|2020-11 | ···················· | ···················· | ···················· | ···················· | ···················· | ···················· |
| Y | 1845 hCoV-19/Poland/79326/2020\|EPI_ISL_732906\|2020-11 | ···················· | ·A·················· | ···················· | ···················· | ···················· | ···················· |
| Y | 1846 hCoV-19/Poland/78563/2020\|EPI_ISL_732907\|2020-11 | ···················· | ····A··············· | ···················· | ···················· | ···················· | ···················· |
| Y | 1847 hCoV-19/Poland/79288/2020\|EPI_ISL_732908\|2020-11 | ···················· | ···················· | ···················· | ···················· | ···················· | ···················· |
| Y | 1848 hCoV-19/Poland/27089/2020\|EPI_ISL_732909\|2020-11 | ···················· | ···················· | ···················· | ···················· | ···················· | ···················· |
| Y | 1849 hCoV-19/Poland/19717/2020\|EPI_ISL_732910\|2020-11 | ···················· | ···················· | ···················· | ···················· | ···················· | ···················· |
| Y | 1850 hCoV-19/Poland/27818/2020\|EPI_ISL_732911\|2020-11 | ···················· | ·········A·········· | ···················· | ···················· | ···················· | ···················· |
| Y | 1851 hCoV-19/Poland/27634/2020\|EPI_ISL_732912\|2020-11 | ···················· | ···················· | ···················· | ···················· | ···················· | ···················· |
| Y | 1852 hCoV-19/Poland/25895/2020\|EPI_ISL_732913\|2020-11 | ···················· | ···················· | ···················· | ···················· | ···················· | ···················· |
| Y | 1853 hCoV-19/Poland/27753/2020\|EPI_ISL_732914\|2020-11 | ···················· | ···················· | ···················· | ···················· | ···················· | ···················· |
| Y | 1854 hCoV-19/Poland/27172/2020\|EPI_ISL_732915\|2020-11 | ···················· | ···················· | ···················· | ···················· | ···················· | ···················· |
| Y | 1855 hCoV-19/Poland/19876/2020\|EPI_ISL_732916\|2020-11 | ···················· | ···················· | ···················· | ···················· | ···················· | ···················· |
| Y | 1856 hCoV-19/Poland/27599/2020\|EPI_ISL_732917\|2020-11 | ···················· | ···················· | ···················· | ···················· | ···················· | ···················· |
| Y | 1857 hCoV-19/Poland/27383/2020\|EPI_ISL_732918\|2020-11 | ···················· | ···················· | ···················· | ···················· | ···················· | ···················· |
| Y | 1858 hCoV-19/Poland/27929/2020\|EPI_ISL_732919\|2020-11 | ···················· | ···················· | ···················· | ···················· | ···················· | ···················· |
| Y | 1859 hCoV-19/Poland/28540/2020\|EPI_ISL_732920\|2020-11 | ···················· | ···················· | ···················· | ···················· | ···················· | ···················· |
| Y | 1861 hCoV-19/Poland/19845/2020\|EPI_ISL_732922\|2020-11 | ···················· | ···················· | ···················· | ···················· | ···················· | ···················· |
| Y | 1862 hCoV-19/Poland/27246/2020\|EPI_ISL_732923\|2020-11 | ···················· | ···················· | ···················· | ···················· | ···················· | ···················· |
| Y | 1863 hCoV-19/Poland/27589/2020\|EPI_ISL_732924\|2020-11 | ···················· | ···················· | ···················· | ···················· | ···················· | ···················· |
| Y | 1864 hCoV-19/Poland/28546/2020\|EPI_ISL_732925\|2020-11 | ···················· | ···················· | ···················· | ···················· | ···················· | ···················· |
| Y | 1865 hCoV-19/Poland/20699/2020\|EPI_ISL_732926\|2020-11 | ···················· | ···················· | ···················· | ···················· | ···················· | ···················· |
| Y | 1866 hCoV-19/mink/Poland/Pomorskie_5/2020\|EPI_ISL_73294 | ···················· | ···················· | ···················· | ···················· | ···················· | ···················· |
| Y | 1867 hCoV-19/mink/Poland/Pomorskie_46/2020\|EPI_ISL_7329 | ···················· | ···················· | ···················· | ···················· | ···················· | ···················· |
| Y | 1868 hCoV-19/mink/Poland/Pomorskie_48/2020\|EPI_ISL_7329 | ···················· | ···················· | ···················· | ···················· | ···················· | ···················· |
| Y | 1869 hCoV-19/mink/Poland/Pomorskie_49/2020\|EPI_ISL_7329 | ···················· | ···················· | ···················· | ···················· | ···················· | ···················· |
| Y | 1870 hCoV-19/mink/Poland/Pomorskie_50/2020\|EPI_ISL_7329 | ···················· | ···················· | ···················· | ···················· | ···················· | ···················· |
| Y | 1871 hCoV-19/mink/Poland/Pomorskie_67/2020\|EPI_ISL_7329 | ···················· | ···················· | ···················· | ···················· | ···················· | ···················· |
| Y | 1872 hCoV-19/mink/Poland/Pomorskie_76/2020\|EPI_ISL_7329 | ···················· | ···················· | ···················· | ···················· | ···················· | ···················· |
| Y | 1873 hCoV-19/mink/Poland/Pomorskie_77/2020\|EPI_ISL_7329 | ···················· | ···················· | ···················· | ···················· | ···················· | ···················· |
| Y | 1874 hCoV-19/mink/Poland/Pomorskie_83/2020\|EPI_ISL_7329 | ···················· | ···················· | ···················· | ···················· | ···················· | ···················· |
| Y | 1875 hCoV-19/mink/Poland/Pomorskie_88/2020\|EPI_ISL_7329 | ···················· | ···················· | ···················· | ···················· | ···················· | ···················· |
| Y | 1876 hCoV-19/Australia/SAP592/2020\|EPI_ISL_732960\|2020- | ···················· | ·A·················· | ···················· | ···················· | ···················· | ··A················· |
| Y | 1877 hCoV-19/New Zealand/20CV0658/2020\|EPI_ISL_732963\|2 | ···················· | ···················· | ···················· | ···················· | ···················· | ···················· |
| Y | 1878 hCoV-19/New Zealand/20CV0662/2020\|EPI_ISL_732964\|2 | ···················· | ···················· | ···················· | ···················· | ···················· | ············G······· |
| Y | 1879 hCoV-19/Belgium/rega-10181080/2020\|EPI_ISL_735120\| | ···················· | ···················· | ···················· | ···················· | ···················· | ···················· |
| Y | 1880 hCoV-19/Belgium/rega-10181081/2020\|EPI_ISL_735121\| | ···················· | ···················· | ···················· | ···················· | ···················· | ···················· |
| Y | 1881 hCoV-19/Belgium/rega-10181082/2020\|EPI_ISL_735122\| | ···················· | ···················· | ···················· | ···················· | ···················· | ···················· |
| Y | 1882 hCoV-19/Belgium/rega-10181083/2020\|EPI_ISL_735123\| | ···················· | ···················· | ···················· | ···················· | ···················· | ···················· |
| Y | 1883 hCoV-19/Belgium/rega-10181084/2020\|EPI_ISL_735124\| | ···················· | ···················· | ···················· | ···················· | ···················· | ···················· |
| Y | 1884 hCoV-19/Belgium/rega-10181085/2020\|EPI_ISL_735125\| | ···················· | ···················· | ···················· | ···················· | ···················· | ···················· |
| Y | 1885 hCoV-19/Belgium/rega-10181086/2020\|EPI_ISL_735126\| | ···················· | ···················· | ···················· | ···················· | ···················· | ···················· |
| Y | 1886 hCoV-19/Belgium/rega-10181087/2020\|EPI_ISL_735127\| | ···················· | ···················· | ···················· | ···················· | ···················· | ···················· |
| Y | 1887 hCoV-19/Belgium/rega-10181088/2020\|EPI_ISL_735128\| | ···················· | ···················· | ···················· | ···················· | ···················· | ···················· |
| Y | 1888 hCoV-19/Belgium/rega-10181089/2020\|EPI_ISL_735129\| | ···················· | ···················· | ···················· | ···················· | ···················· | ···················· |
| Y | 1889 hCoV-19/Belgium/rega-10181090/2020\|EPI_ISL_735130\| | ···················· | ·········A·········· | ···················· | ···················· | ···················· | ···················· |
| Y | 1890 hCoV-19/Belgium/rega-10181091/2020\|EPI_ISL_735131\| | ···················· | ···················· | ···················· | ···················· | ···················· | ···················· |
| Y | 1891 hCoV-19/Belgium/rega-10181111/2020\|EPI_ISL_735132\| | ···················· | ···················· | ···················· | ···················· | ···················· | ···················· |
| Y | 1892 hCoV-19/Belgium/rega-10181112/2020\|EPI_ISL_735133\| | ···················· | ···················· | ···················· | ···················· | ···················· | ···················· |
| Y | 1893 hCoV-19/Belgium/rega-10181113/2020\|EPI_ISL_735134\| | ···················· | ···················· | ···················· | ···················· | ···················· | ···················· |
| Y | 1894 hCoV-19/Belgium/rega-10181163/2020\|EPI_ISL_735135\| | ···················· | ···················· | ···················· | ···················· | ···················· | ···················· |
| Y | 1895 hCoV-19/Belgium/rega-10181164/2020\|EPI_ISL_735136\| | ···················· | ···················· | ···················· | ···················· | ···················· | ···················· |
| Y | 1896 hCoV-19/Belgium/rega-10181165/2020\|EPI_ISL_735137\| | ···················· | ···················· | ···················· | ···················· | ···················· | ···················· |
| Y | 1897 hCoV-19/Belgium/rega-10181166/2020\|EPI_ISL_735138\| | ···················· | ···················· | ···················· | ···················· | ···················· | ···················· |
| Y | 1898 hCoV-19/Belgium/rega-10181167/2020\|EPI_ISL_735139\| | ···················· | ···················· | ···················· | ···················· | ···················· | ···················· |
| Y | 1899 hCoV-19/Belgium/rega-10191092/2020\|EPI_ISL_735140\| | ···················· | ···················· | ···················· | ···················· | ···················· | ···················· |
| Y | 1900 hCoV-19/Belgium/rega-10191093/2020\|EPI_ISL_735141\| | ···················· | ···················· | ···················· | ···················· | ···················· | ···················· |
| Y | 1901 hCoV-19/Belgium/rega-10191094/2020\|EPI_ISL_735142\| | ···················· | ···················· | ···················· | ···················· | ···················· | ···················· |
| Y | 1902 hCoV-19/Belgium/rega-10191095/2020\|EPI_ISL_735143\| | ···················· | ···················· | ···················· | ···················· | ···················· | ···················· |

| | | | | | | | | |
|---|---|---|---|---|---|---|---|---|
| Y | 1903 | hCoV-19/Belgium/rega-10191097/2020\|EPI_ISL_735144\| | ···················· | ···················· | ···················· | ···················· | ···················· | ···················· |
| Y | 1904 | hCoV-19/Belgium/rega-10191098/2020\|EPI_ISL_735145\| | ···················· | ···················· | ···················· | ···················· | ···················· | ···················· |
| Y | 1905 | hCoV-19/Belgium/rega-10191099/2020\|EPI_ISL_735146\| | ···················· | ···················· | ···················· | ···················· | ···················· | ···················· |
| Y | 1906 | hCoV-19/Belgium/rega-10191100/2020\|EPI_ISL_735147\| | ···················· | ···················· | ···················· | ···················· | ···················· | ···················· |
| Y | 1907 | hCoV-19/Belgium/rega-10191101/2020\|EPI_ISL_735148\| | ···················· | ···················· | ···················· | ···················· | ···················· | ···················· |
| Y | 1908 | hCoV-19/Belgium/rega-10191102/2020\|EPI_ISL_735149\| | ···················· | ···················· | ···················· | ···················· | ···················· | ···················· |
| Y | 1909 | hCoV-19/Belgium/rega-10191129/2020\|EPI_ISL_735160\| | ···················· | ···················· | ···················· | ···················· | ···················· | ···················· |
| Y | 1910 | hCoV-19/Belgium/rega-10191130/2020\|EPI_ISL_735161\| | ···················· | ···················· | ···················· | ···················· | ···················· | ···················· |
| Y | 1911 | hCoV-19/Belgium/rega-10191131/2020\|EPI_ISL_735162\| | ···················· | ···················· | ···················· | ···················· | ···················· | ···················· |
| Y | 1912 | hCoV-19/Belgium/rega-10191222/2020\|EPI_ISL_735163\| | ···················· | ···················· | ···················· | ···················· | ···················· | ···················· |
| Y | 1913 | hCoV-19/Belgium/rega-10191228/2020\|EPI_ISL_735164\| | ···················· | ···················· | ···················· | ···················· | ···················· | ···················· |
| Y | 1914 | hCoV-19/Belgium/rega-10191234/2020\|EPI_ISL_735165\| | ···················· | ···················· | ···················· | ···················· | ···················· | ···················· |
| Y | 1916 | hCoV-19/Belgium/rega-10191246/2020\|EPI_ISL_735167\| | ···················· | ···················· | ···················· | ···················· | ···················· | ···················· |
| Y | 1917 | hCoV-19/Belgium/rega-10191252/2020\|EPI_ISL_735168\| | ···················· | ···················· | ···················· | ···················· | ···················· | ···················· |
| Y | 1918 | hCoV-19/Belgium/rega-10191258/2020\|EPI_ISL_735169\| | ···················· | ···················· | ···················· | ···················· | ···················· | ···················· |
| Y | 1919 | hCoV-19/Belgium/rega-10191259/2020\|EPI_ISL_735170\| | ···················· | ···················· | ···················· | ···················· | ···················· | ···················· |
| Y | 1920 | hCoV-19/Belgium/rega-10191260/2020\|EPI_ISL_735171\| | ···················· | ···················· | ···················· | ···················· | ···················· | ···················· |
| Y | 1921 | hCoV-19/Belgium/rega-10191262/2020\|EPI_ISL_735172\| | ···················· | ···················· | ···················· | ···················· | ···················· | ···················· |
| Y | 1922 | hCoV-19/Belgium/rega-10191263/2020\|EPI_ISL_735173\| | ···················· | ···················· | ···················· | ···················· | ···················· | ···················· |
| Y | 1923 | hCoV-19/Belgium/rega-10191264/2020\|EPI_ISL_735174\| | ···················· | ···················· | ···················· | ···················· | ···················· | ···················· |
| Y | 1924 | hCoV-19/Belgium/rega-10191265/2020\|EPI_ISL_735175\| | ···················· | ···················· | ···················· | ···················· | ···················· | ···················· |
| Y | 1925 | hCoV-19/Belgium/rega-10191266/2020\|EPI_ISL_735176\| | ···················· | ···················· | ···················· | ···················· | ···················· | ···················· |
| Y | 1926 | hCoV-19/Belgium/rega-10191267/2020\|EPI_ISL_735177\| | ···················· | ···················· | ···················· | ···················· | ···················· | ···················· |
| Y | 1927 | hCoV-19/Belgium/rega-10191268/2020\|EPI_ISL_735178\| | ···················· | ···················· | ···················· | ···················· | ···················· | ···················· |
| Y | 1928 | hCoV-19/Belgium/rega-10191269/2020\|EPI_ISL_735179\| | ···················· | ···················· | ···················· | ···················· | ···················· | ···················· |
| Y | 1929 | hCoV-19/Belgium/rega-10191270/2020\|EPI_ISL_735180\| | ···················· | ···················· | ···················· | ···················· | ···················· | ···················· |
| Y | 1930 | hCoV-19/Belgium/rega-10191271/2020\|EPI_ISL_735181\| | ···················· | ···················· | ···················· | ···················· | ···················· | ···················· |
| Y | 1931 | hCoV-19/Belgium/rega-10191272/2020\|EPI_ISL_735182\| | ···················· | ···················· | ···················· | ···················· | ···················· | ···················· |
| Y | 1932 | hCoV-19/Belgium/rega-10191274/2020\|EPI_ISL_735183\| | ···················· | ···················· | ···················· | ···················· | ···················· | ···················· |
| Y | 1933 | hCoV-19/Belgium/rega-10191275/2020\|EPI_ISL_735184\| | ···················· | ···················· | ···················· | ···················· | ···················· | ···················· |
| Y | 1934 | hCoV-19/Belgium/rega-10191276/2020\|EPI_ISL_735185\| | ···················· | ···················· | ···················· | ···················· | ···················· | ···················· |
| Y | 1935 | hCoV-19/Belgium/rega-10191278/2020\|EPI_ISL_735186\| | ···················· | ···················· | ···················· | ···················· | ···················· | ···················· |
| Y | 1936 | hCoV-19/Belgium/rega-10191279/2020\|EPI_ISL_735187\| | ···················· | ···················· | ···················· | ···················· | ···················· | ···················· |
| Y | 1937 | hCoV-19/Belgium/rega-10191280/2020\|EPI_ISL_735188\| | ···················· | ···················· | ···················· | ···················· | ···················· | ···················· |
| Y | 1938 | hCoV-19/Belgium/rega-10191282/2020\|EPI_ISL_735189\| | ···················· | ···················· | ···················· | ···················· | ···················· | ···················· |
| Y | 1939 | hCoV-19/Belgium/rega-10191283/2020\|EPI_ISL_735190\| | ···················· | ···················· | ···················· | ···················· | ···················· | ···················· |
| Y | 1940 | hCoV-19/Belgium/rega-10191284/2020\|EPI_ISL_735191\| | ···················· | ···················· | ···················· | ···················· | ···················· | ···················· |
| Y | 1941 | hCoV-19/Belgium/rega-10191285/2020\|EPI_ISL_735192\| | ···················· | ···················· | ···················· | ···················· | ···················· | ···················· |
| Y | 1942 | hCoV-19/Belgium/rega-10191286/2020\|EPI_ISL_735193\| | ···················· | ···················· | ···················· | ···················· | ···················· | ···················· |
| Y | 1943 | hCoV-19/Belgium/rega-10191288/2020\|EPI_ISL_735194\| | ···················· | ···················· | ···················· | ···················· | ···················· | ···················· |
| Y | 1945 | hCoV-19/Belgium/rega-10191291/2020\|EPI_ISL_735196\| | ···················· | ···················· | ···················· | ···················· | ···················· | ···················· |
| Y | 1946 | hCoV-19/Belgium/rega-10191292/2020\|EPI_ISL_735197\| | ···················· | ···················· | ···················· | ···················· | ···················· | ···················· |
| Y | 1947 | hCoV-19/Belgium/rega-10191293/2020\|EPI_ISL_735198\| | ···················· | ···················· | ···················· | ···················· | ···················· | ···················· |
| Y | 1948 | hCoV-19/Belgium/rega-10191294/2020\|EPI_ISL_735199\| | ···················· | ···················· | ···················· | ···················· | ···················· | ···················· |
| Y | 1949 | hCoV-19/England/MILK-C52A05/2020\|EPI_ISL_736452\|20 | ···················· | ···················· | ···················· | ···················· | ···················· | ···················· |
| Y | 1950 | hCoV-19/England/QEUH-C53EEA/2020\|EPI_ISL_736455\|20 | ···················· | ···················· | ···················· | ···················· | ···················· | ···················· |
| Y | 1951 | hCoV-19/England/QEUH-C53FF6/2020\|EPI_ISL_736456\|20 | ···················· | ···················· | ···················· | ···················· | ···················· | ···················· |
| Y | 1952 | hCoV-19/England/QEUH-C54502/2020\|EPI_ISL_736461\|20 | ···················· | ···················· | ···················· | ···················· | ···················· | ···················· |
| Y | 1953 | hCoV-19/England/MILK-C52CC3/2020\|EPI_ISL_736463\|20 | ···················· | ···················· | ···················· | ···················· | ···················· | ···················· |
| Y | 1954 | hCoV-19/England/QEUH-C53E53/2020\|EPI_ISL_736464\|20 | ···················· | ···················· | ···················· | ···················· | ···················· | ···················· |
| Y | 1955 | hCoV-19/England/MILK-C52A7E/2020\|EPI_ISL_736467\|20 | ···················· | ···················· | ···················· | ···················· | ···················· | ···················· |
| Y | 1956 | hCoV-19/England/QEUH-C5799A/2020\|EPI_ISL_736470\|20 | ···················· | ···················· | ···················· | ···················· | ···················· | ···················· |
| Y | 1957 | hCoV-19/England/QEUH-C541A4/2020\|EPI_ISL_736471\|20 | ···················· | ···················· | ···················· | ···················· | ···················· | ···················· |
| Y | 1958 | hCoV-19/England/QEUH-C545C6/2020\|EPI_ISL_736478\|20 | ···················· | ···················· | ···················· | ···················· | ···················· | ···················· |
| Y | 1959 | hCoV-19/England/QEUH-C4BE5E/2020\|EPI_ISL_736492\|20 | ···················· | ···················· | ···················· | ···················· | ···················· | ···················· |
| Y | 1960 | hCoV-19/England/ALDP-C48817/2020\|EPI_ISL_736494\|20 | ···················· | ···················· | ···················· | ···················· | ···················· | ···················· |
| Y | 1961 | hCoV-19/England/ALDP-C48B0F/2020\|EPI_ISL_736496\|20 | ···················· | ···················· | ···················· | ···················· | ···················· | ···················· |
| Y | 1962 | hCoV-19/England/MILK-C50472/2020\|EPI_ISL_736497\|20 | ···················· | ·A·················· | ···················· | ···················· | ···················· | ··A················· |
| Y | 1963 | hCoV-19/England/ALDP-C485B6/2020\|EPI_ISL_736498\|20 | ···················· | ···················· | ···················· | ···················· | ···················· | ···················· |
| Y | 1964 | hCoV-19/Switzerland/SH-ETHZ-410733/2020\|EPI_ISL_73 | ···················· | ···················· | ···················· | ···················· | ···················· | ···················· |
| Y | 1965 | hCoV-19/Switzerland/SH-ETHZ-410734/2020\|EPI_ISL_73 | ···················· | ···················· | ···················· | ···················· | ···············K···· | ···················· |
| Y | 1966 | hCoV-19/Switzerland/SZ-ETHZ-410043/2020\|EPI_ISL_73 | ···················· | ···················· | ···················· | ···················· | ···················· | ···················· |
| Y | 1967 | hCoV-19/Switzerland/SZ-ETHZ-410194/2020\|EPI_ISL_73 | ···················· | ···················· | ···················· | ···················· | ···················· | ···················· |
| Y | 1968 | hCoV-19/Switzerland/SO-ETHZ-320193/2020\|EPI_ISL_73 | ···················· | ···················· | ···················· | ···················· | ···················· | ···················· |
| Y | 1969 | hCoV-19/Switzerland/SO-ETHZ-410039/2020\|EPI_ISL_73 | ···················· | ···················· | ···················· | ···················· | ···················· | ···················· |
| Y | 1970 | hCoV-19/Switzerland/SO-ETHZ-410048/2020\|EPI_ISL_73 | ···················· | ···················· | ···················· | ···················· | ···················· | ···················· |
| Y | 1971 | hCoV-19/Switzerland/SO-ETHZ-410058/2020\|EPI_ISL_73 | ···················· | ···················· | ···················· | ···················· | ···················· | ···················· |
| Y | 1972 | hCoV-19/Switzerland/SO-ETHZ-410059/2020\|EPI_ISL_73 | ···················· | ···················· | ···················· | ···················· | ···················· | ···················· |
| Y | 1973 | hCoV-19/Switzerland/SO-ETHZ-410060/2020\|EPI_ISL_73 | ···················· | ···················· | ···················· | ···················· | ···················· | ···················· |
| Y | 1974 | hCoV-19/Switzerland/SO-ETHZ-410061/2020\|EPI_ISL_73 | ···················· | ···················· | ···················· | ···················· | ···················· | ···················· |
| Y | 1975 | hCoV-19/Switzerland/SO-ETHZ-410071/2020\|EPI_ISL_73 | ···················· | ···················· | ···················· | ···················· | ···················· | ···················· |
| Y | 1976 | hCoV-19/Switzerland/SO-ETHZ-410122/2020\|EPI_ISL_73 | ···················· | ···················· | ···················· | ···················· | ···················· | ···················· |
| Y | 1977 | hCoV-19/Switzerland/SO-ETHZ-410129/2020\|EPI_ISL_73 | ···················· | ···················· | ···················· | ···················· | ···················· | ···················· |
| Y | 1979 | hCoV-19/Switzerland/SO-ETHZ-410237/2020\|EPI_ISL_73 | ········Y··········· | ···················· | ···················· | ···················· | ···················· | ···················· |
| Y | 1980 | hCoV-19/Switzerland/SO-ETHZ-410290/2020\|EPI_ISL_73 | ···················· | ···················· | ···················· | ···················· | ···················· | ···················· |
| Y | 1981 | hCoV-19/Switzerland/SO-ETHZ-410302/2020\|EPI_ISL_73 | ···················· | ···················· | ···················· | ···················· | ···················· | ···················· |
| Y | 1982 | hCoV-19/Switzerland/SO-ETHZ-410338/2020\|EPI_ISL_73 | ···················· | ······A············· | ···················· | ···················· | ···················· | ···················· |
| Y | 1983 | hCoV-19/Switzerland/SO-ETHZ-410345/2020\|EPI_ISL_73 | ···················· | ···················· | ···················· | ···················· | ···················· | ···················· |
| Y | 1984 | hCoV-19/USA/MS-CDC-2-3693659/2020\|EPI_ISL_747100\|2 | ···················· | ···················· | ···················· | ···················· | ···················· | ···················· |
| Y | 1985 | hCoV-19/USA/MS-CDC-2-3693783/2020\|EPI_ISL_747101\|2 | ···················· | ·············A······ | ···················· | ···················· | ···················· | ···················· |

```
Y 1986 hCoV-19/USA/MS-CDC-2-3693768/2020|EPI_ISL_747102|2 ···················· ···················· ····················· ···················· ···················· ····················
Y 1987 hCoV-19/USA/IL-CDC-2-3693758/2020|EPI_ISL_747103|2 ···················· ···················· ····················· ···················· ···················· ····················
Y 1988 hCoV-19/USA/IL-CDC-2-3693777/2020|EPI_ISL_747104|2 ···················· ···················· ····················· ···················· ···················· ····················
Y 1989 hCoV-19/USA/IL-CDC-2-3693786/2020|EPI_ISL_747105|2 ···················· ···················· ····················· ···················· ···················· ····················
Y 1990 hCoV-19/USA/IL-CDC-2-3693680/2020|EPI_ISL_747106|2 ···················· ···················· ····················· ···················· ·T·················· ····················
Y 1991 hCoV-19/USA/NJ-CDC-2-3693780/2020|EPI_ISL_747107|2 ···················· ···················· ····················· ···················· ···················· ····················
Y 1992 hCoV-19/USA/NJ-CDC-2-3693666/2020|EPI_ISL_747108|2 ···················· ···················· ····················· ···················· ···················· ····················
Y 1993 hCoV-19/USA/NJ-CDC-2-3693764/2020|EPI_ISL_747110|2 ···················· ···················· ····················· ···················· ···················· ····················
Y 1994 hCoV-19/USA/NJ-CDC-2-3693474/2020|EPI_ISL_747111|2 ···················· ···················· ····················· ···················· ···················· ····················
Y 1995 hCoV-19/USA/MI-CDC-2-3693689/2020|EPI_ISL_747112|2 ···················· ···················· ····················· ···················· ···················· ····················
Y 1996 hCoV-19/USA/MI-CDC-2-3693663/2020|EPI_ISL_747113|2 ···················· ·············A······ ····················· ···················· ···················· ····················
Y 1997 hCoV-19/USA/MI-CDC-2-3693788/2020|EPI_ISL_747114|2 ···················· ···················· ····················· ···················· ···················· ····················
Y 1998 hCoV-19/USA/MI-CDC-2-3693684/2020|EPI_ISL_747116|2 ···················· ·············M······ ····················· ···················· ···················· ····················
Y 1999 hCoV-19/USA/NC-CDC-2-3693685/2020|EPI_ISL_747117|2 ···················· ···················· ····················· ···················· ···················· ····················
Y 2000 hCoV-19/USA/NC-CDC-2-3693695/2020|EPI_ISL_747118|2 ···················· ·A·················· ····················· ···················· ···················· ····················
Y 2001 hCoV-19/USA/NC-CDC-2-3693555/2020|EPI_ISL_747119|2 ···················· ···················· ····················· ···················· ···················· ····················
Y 2002 hCoV-19/USA/KS-CDC-2-3693581/2020|EPI_ISL_747140|2 ···················· ···················· ····················· ···················· ···················· ····················
Y 2003 hCoV-19/USA/MO-CDC-2-3693556/2020|EPI_ISL_747141|2 ···················· ·R·················· ····················· ···················· ···················· ····················
Y 2004 hCoV-19/USA/MO-CDC-2-3693463/2020|EPI_ISL_747142|2 ···················· ···················· ····················· ···················· ···················· ····················
Y 2005 hCoV-19/USA/MO-CDC-2-3693485/2020|EPI_ISL_747143|2 ···················· ···················· ····················· ···················· ···················· ····················
Y 2006 hCoV-19/USA/MO-CDC-2-3693577/2020|EPI_ISL_747144|2 ···················· ···················· ····················· ···················· ···················· ····················
Y 2007 hCoV-19/USA/MO-CDC-2-3693582/2020|EPI_ISL_747145|2 ···················· ···················· ····················· ···················· ···················· ····················
Y 2008 hCoV-19/USA/TX-CDC-2-3693566/2020|EPI_ISL_747146|2 ···················· ···················· ····················· ···················· ···················· ····················
Y 2009 hCoV-19/USA/TX-CDC-2-3693585/2020|EPI_ISL_747147|2 ···················· ···················· ····················· ···················· ···················· ····················
Y 2010 hCoV-19/USA/TX-CDC-2-3693470/2020|EPI_ISL_747149|2 ···················· ···················· ····················· ···················· ···················· ····················
Y 2011 hCoV-19/USA/TX-CDC-2-3693478/2020|EPI_ISL_747150|2 ···················· ·R·················· ····················· ···················· ···················· ····················
Y 2012 hCoV-19/USA/NV-CDC-2-3693483/2020|EPI_ISL_747151|2 ···················· ·A·················· ····················· ···················· ···················· ····················
Y 2013 hCoV-19/USA/NV-CDC-2-3693465/2020|EPI_ISL_747152|2 ···················· ···················· ····················· ···················· ···················· ····················
Y 2014 hCoV-19/USA/NV-CDC-2-3693578/2020|EPI_ISL_747153|2 ···················· ·R·················· ····················· ···················· ···················· ····················
Y 2015 hCoV-19/USA/NV-CDC-2-3693480/2020|EPI_ISL_747154|2 ···················· ·R·················· ····················· ···················· ···················· ····················
Y 2016 hCoV-19/USA/OK-CDC-2-3693599/2020|EPI_ISL_747155|2 ········T··········· ·R·················· ····················· ···················· ···················· ····················
Y 2017 hCoV-19/USA/OK-CDC-2-3693488/2020|EPI_ISL_747156|2 ···················· ·R·················· ····················· ···················· ···················· ····················
Y 2018 hCoV-19/USA/OK-CDC-2-3693595/2020|EPI_ISL_747157|2 ···················· ···················· ····················· ···················· ···················· ····················
Y 2019 hCoV-19/USA/OK-CDC-2-3693479/2020|EPI_ISL_747158|2 ···················· ·R·················· ····················· ···················· ···················· ····················
Y 2020 hCoV-19/USA/OK-CDC-2-3693466/2020|EPI_ISL_747160|2 ···················· ·R·················· ····················· ···················· ···················· ····················
Y 2021 hCoV-19/USA/OK-CDC-2-3693469/2020|EPI_ISL_747161|2 ···················· ···················· ····················· ···················· ···················· ····················
Y 2022 hCoV-19/USA/OK-CDC-2-3693590/2020|EPI_ISL_747162|2 ···················· ···················· ····················· ···················· ···················· ····················
Y 2023 hCoV-19/USA/OK-CDC-2-3693462/2020|EPI_ISL_747163|2 ···················· ···················· ····················· ···················· ···················· ····················
Y 2024 hCoV-19/USA/OR-CDC-2-3693558/2020|EPI_ISL_747166|2 ···················· ·A·················· ····················· ···················· ···················· ····················
Y 2025 hCoV-19/USA/OR-CDC-2-3693579/2020|EPI_ISL_747167|2 ···················· ···················· ····················· ···················· ···················· ····················
Y 2026 hCoV-19/USA/OR-CDC-2-3693560/2020|EPI_ISL_747168|2 ···················· ···················· ····················· ···················· ···················· ····················
Y 2027 hCoV-19/USA/OR-CDC-2-3693487/2020|EPI_ISL_747169|2 ···················· ·R·················· ····················· ···················· ···················· ····················
Y 2028 hCoV-19/USA/OR-CDC-2-3693481/2020|EPI_ISL_747170|2 ···················· ···················· ····················· ···················· ···················· ····················
Y 2029 hCoV-19/USA/OR-CDC-2-3693468/2020|EPI_ISL_747171|2 ···················· ···················· ····················· ···················· ···················· ····················
Y 2030 hCoV-19/USA/OR-CDC-2-3693583/2020|EPI_ISL_747172|2 ···················· ···················· ····················· ···················· ···················· ····················
Y 2031 hCoV-19/USA/NC-CDC-2-4693552/2020|EPI_ISL_747175|2 ···················· ···················· ····················· ···················· ···················· ····················
Y 2032 hCoV-19/USA/NC-CDC-2-4693563/2020|EPI_ISL_747176|2 ···················· ···················· ····················· ···················· ···················· ····················
Y 2033 hCoV-19/USA/NC-CDC-2-4693589/2020|EPI_ISL_747177|2 ···················· ···················· ····················· ···················· ···················· ····················
Y 2035 hCoV-19/USA/NC-CDC-2-3693561/2020|EPI_ISL_747179|2 ···················· ···················· ····················· ···················· ···················· ····················
Y 2036 hCoV-19/USA/NC-CDC-2-3693473/2020|EPI_ISL_747180|2 ···················· ···················· ····················· ···················· ···················· ····················
Y 2037 hCoV-19/USA/NC-CDC-2-3693467/2020|EPI_ISL_747182|2 ···················· ···················· ····················· ···················· ···················· ····················
Y 2038 hCoV-19/USA/TN-CDC-2-3693573/2020|EPI_ISL_747183|2 ···················· ···················· ····················· ···················· ···················· ····················
Y 2039 hCoV-19/USA/TN-CDC-2-3693597/2020|EPI_ISL_747184|2 ···················· ···················· ····················· ···················· ···················· ····················
Y 2040 hCoV-19/USA/TN-CDC-2-3693460/2020|EPI_ISL_747185|2 ···················· ·············A······ ····················· ···················· ···················· ····················
Y 2041 hCoV-19/USA/TN-CDC-2-3693461/2020|EPI_ISL_747186|2 ···················· ·R·················· ····················· ···················· ···················· ····················
Y 2042 hCoV-19/USA/TN-CDC-2-3693571/2020|EPI_ISL_747187|2 ···················· ···················· ····················· ···················· ···················· ····················
Y 2043 hCoV-19/USA/TN-CDC-2-3693587/2020|EPI_ISL_747188|2 ···················· ···················· ····················· ···················· ···················· ····················
Y 2044 hCoV-19/USA/OR-CDC-2-3693562/2020|EPI_ISL_747189|2 ···················· ···················· ····················· ···················· ···················· ····················
Y 2045 hCoV-19/USA/OR-CDC-2-3693471/2020|EPI_ISL_747190|2 ···················· ·R·················· ····················· ···················· ···················· ····················
Y 2046 hCoV-19/USA/OR-CDC-2-3693497/2020|EPI_ISL_747191|2 ···················· ···················· ····················· ···················· ···················· ····················
Y 2047 hCoV-19/USA/OR-CDC-2-3693600/2020|EPI_ISL_747192|2 ···················· ·R·················· ····················· ···················· ···················· ····················
Y 2048 hCoV-19/USA/OR-CDC-2-3693491/2020|EPI_ISL_747193|2 ···················· ···················· ····················· ···················· ···················· ····················
Y 2049 hCoV-19/England/LIVE-DCC3CE/2020|EPI_ISL_724502|20 ···················· ···················· ····················· ···················· ···················· ····················
Y 2050 hCoV-19/England/LIVE-DCC3EC/2020|EPI_ISL_724503|20 ···················· ···················· ····················· ···················· ···················· ····················
Y 2051 hCoV-19/Wales/PHWC-492F8A/2020|EPI_ISL_725815|2020 ···················· ···················· ····················· ···················· ···················· ····················
Y 2052 hCoV-19/Wales/PHWC-492F99/2020|EPI_ISL_725816|2020 ···················· ···················· ····················· ···················· ···················· ····················
Y 2053 hCoV-19/Wales/PHWC-492FA8/2020|EPI_ISL_725817|2020 ···················· ···················· ····················· ···················· ···················· ····················
Y 2054 hCoV-19/Wales/PHWC-492FB7/2020|EPI_ISL_725818|2020 ···················· ···················· ····················· ···················· ···················· ····················
Y 2055 hCoV-19/Wales/PHWC-492FC6/2020|EPI_ISL_725819|2020 ···················· ···················· ····················· ···················· ···················· ····················
Y 2056 hCoV-19/Wales/PHWC-492FD5/2020|EPI_ISL_725820|2020 ···················· ···················· ····················· ···················· ···················· ····················
Y 2057 hCoV-19/Wales/PHWC-492FE4/2020|EPI_ISL_725821|2020 ···················· ···················· ····················· ···················· ···················· ····················
Y 2058 hCoV-19/Wales/PHWC-492FF3/2020|EPI_ISL_725822|2020 ···················· ···················· ····················· ···················· ···················· ····················
Y 2061 hCoV-19/Wales/PHWC-49302C/2020|EPI_ISL_725825|2020 ···················· ···················· ····················· ···················· ···················· ····················
Y 2062 hCoV-19/Wales/PHWC-4930A4/2020|EPI_ISL_725833|2020 ···················· ···················· ····················· ···················· ···················· ····················
Y 2063 hCoV-19/Wales/PHWC-4930B3/2020|EPI_ISL_725834|2020 ···················· ···················· ····················· ···················· ···················· ····················
Y 2064 hCoV-19/Wales/PHWC-4930C2/2020|EPI_ISL_725835|2020 ···················· ···················· ····················· ···················· ···················· ····················
Y 2065 hCoV-19/Wales/PHWC-4930D1/2020|EPI_ISL_725836|2020 ···················· ···················· ····················· ···················· ···················· ····················
Y 2066 hCoV-19/Japan/FHU-CS56-0221/2020|EPI_ISL_728000|20 ···················· ···················· ····················· ···················· ···················· ····················
Y 2067 hCoV-19/USA/WI-UW-2231/2020|EPI_ISL_728001|2020-11 ···················· ···················· ····················· ···················· ···················· ····················
Y 2069 hCoV-19/USA/WI-UW-2234/2020|EPI_ISL_728004|2020-11 ···················· ···················· ····················· ···················· ···················· ····················
```

| | | | | | | | | |
|---|---|---|---|---|---|---|---|---|
| Y | 2070 | hCoV-19/USA/WI-UW-2235/2020\|EPI_ISL_728005\|2020-11 | ···················· | ···················· | ···················· | ···················· | ···················· | ···················· |
| Y | 2071 | hCoV-19/USA/WI-UW-2236/2020\|EPI_ISL_728006\|2020-11 | ···················· | ···················· | ···················· | ···················· | ···················· | ···················· |
| Y | 2072 | hCoV-19/USA/IL-UW-2238/2020\|EPI_ISL_728007\|2020-11 | ···················· | ···················· | ···················· | ···················· | ···················· | ···················· |
| Y | 2073 | hCoV-19/USA/WI-UW-2239/2020\|EPI_ISL_728008\|2020-11 | ···················· | ···················· | ···················· | ···················· | ···················· | ···················· |
| Y | 2075 | hCoV-19/USA/WI-UW-2241/2020\|EPI_ISL_728010\|2020-11 | ···················· | ···················· | ···················· | ···················· | ···················· | ···················· |
| Y | 2076 | hCoV-19/USA/WI-UW-2242/2020\|EPI_ISL_728011\|2020-11 | ···················· | ···················· | ···················· | ···················· | ···················· | ···················· |
| Y | 2078 | hCoV-19/USA/WI-UW-2244/2020\|EPI_ISL_728013\|2020-09 | ···················· | ···················· | ···················· | ···················· | ···················· | ···················· |
| Y | 2079 | hCoV-19/USA/WI-UW-2245/2020\|EPI_ISL_728014\|2020-10 | ···················· | ···················· | ···················· | ···················· | ···················· | ···················· |
| Y | 2080 | hCoV-19/USA/IL-UW-2246/2020\|EPI_ISL_728015\|2020-10 | ···················· | ···················· | ···················· | ···················· | ···················· | ···················· |
| Y | 2081 | hCoV-19/USA/WI-UW-2247/2020\|EPI_ISL_728016\|2020-09 | ···················· | ···················· | ···················· | ···················· | ···················· | ···················· |
| Y | 2084 | hCoV-19/USA/WI-UW-2252/2020\|EPI_ISL_728021\|2020-10 | ···················· | ···················· | ···················· | ···················· | ···················· | ···················· |
| Y | 2085 | hCoV-19/USA/WI-UW-2253/2020\|EPI_ISL_728022\|2020-09 | ···················· | ···················· | ···················· | ···················· | ···················· | ···················· |
| Y | 2086 | hCoV-19/USA/WI-UW-2254/2020\|EPI_ISL_728023\|2020-10 | ···················· | ···················· | ···················· | ···················· | ···················· | ···················· |
| Y | 2088 | hCoV-19/USA/IL-UW-2256/2020\|EPI_ISL_728025\|2020-09 | ···················· | ·A·················· | ···················· | ···················· | ···················· | ···················· |
| Y | 2089 | hCoV-19/USA/WI-UW-2257/2020\|EPI_ISL_728026\|2020-09 | ···················· | ···················· | ···················· | ···················· | ···················· | ···················· |
| Y | 2090 | hCoV-19/USA/IL-UW-2258/2020\|EPI_ISL_728027\|2020-10 | ···················· | ···················· | ···················· | ···················· | ···················· | ···················· |
| Y | 2091 | hCoV-19/USA/WI-UW-2259/2020\|EPI_ISL_728028\|2020-10 | ···················· | ···················· | ···················· | ···················· | ···················· | ···················· |
| Y | 2092 | hCoV-19/USA/WI-UW-2261/2020\|EPI_ISL_728030\|2020-10 | ···················· | ···················· | ···················· | ···················· | ···················· | ···················· |
| Y | 2093 | hCoV-19/USA/WI-UW-2262/2020\|EPI_ISL_728031\|2020-10 | ···················· | ···················· | ···················· | ···················· | ···················· | ···················· |
| Y | 2094 | hCoV-19/USA/WI-UW-2263/2020\|EPI_ISL_728032\|2020-10 | ···················· | ···················· | ···················· | ···················· | ···················· | ···················· |
| Y | 2095 | hCoV-19/USA/WI-UW-2264/2020\|EPI_ISL_728033\|2020-10 | ···················· | ···················· | ···················· | ···················· | ···················· | ···················· |
| Y | 2096 | hCoV-19/USA/WI-UW-2265/2020\|EPI_ISL_728034\|2020-11 | ···················· | ···················· | ···················· | ···················· | ···················· | ···················· |
| Y | 2097 | hCoV-19/USA/WI-UW-2266/2020\|EPI_ISL_728035\|2020-10 | ···················· | ···················· | ···················· | ···················· | ···················· | ···················· |
| Y | 2101 | hCoV-19/USA/WI-UW-2270/2020\|EPI_ISL_728039\|2020-11 | ···················· | ···················· | ···················· | ···················· | ···················· | ···················· |
| Y | 2103 | hCoV-19/USA/WI-UW-2273/2020\|EPI_ISL_728041\|2020-10 | ···················· | ···················· | ···················· | ···················· | ···················· | ···················· |
| Y | 2106 | hCoV-19/USA/WI-UW-2277/2020\|EPI_ISL_728044\|2020-11 | ···················· | ···················· | ···················· | ···················· | ···················· | ···················· |
| Y | 2108 | hCoV-19/USA/WI-UW-2279/2020\|EPI_ISL_728046\|2020-11 | ···················· | ···················· | ···················· | ···················· | ···················· | ···················· |
| Y | 2117 | hCoV-19/USA/WI-UW-2289/2020\|EPI_ISL_728055\|2020-11 | ···················· | ···················· | ···················· | ···················· | ···················· | ···················· |
| Y | 2132 | hCoV-19/USA/WI-UW-2324/2020\|EPI_ISL_728090\|2020-11 | ···················· | ···················· | ···················· | ···················· | ···················· | ···················· |
| Y | 2133 | hCoV-19/USA/WI-UW-2325/2020\|EPI_ISL_728091\|2020-11 | ···················· | ···················· | ···················· | ···················· | ···················· | ···················· |
| Y | 2134 | hCoV-19/USA/WI-UW-2326/2020\|EPI_ISL_728092\|2020-10 | ···················· | ···················· | ···················· | ···················· | ···················· | ···················· |
| Y | 2135 | hCoV-19/USA/IL-UW-2327/2020\|EPI_ISL_728093\|2020-09 | ···················· | ·A·················· | ···················· | ···················· | ···················· | ···················· |
| Y | 2136 | hCoV-19/USA/WI-UW-2328/2020\|EPI_ISL_728094\|2020-11 | ···················· | ···················· | ···················· | ···················· | ···················· | ···················· |
| Y | 2137 | hCoV-19/USA/WI-UW-2329/2020\|EPI_ISL_728095\|2020-11 | ···················· | ···················· | ···················· | ···················· | ···················· | ···················· |
| Y | 2139 | hCoV-19/USA/WI-UW-2331/2020\|EPI_ISL_728097\|2020-11 | ···················· | ···················· | ···················· | ···················· | ···················· | ···················· |
| Y | 2140 | hCoV-19/USA/WI-UW-2332/2020\|EPI_ISL_728098\|2020-11 | ···················· | ···················· | ···················· | ···················· | ···················· | ···················· |
| Y | 2141 | hCoV-19/Canada/ON-S1904/2020\|EPI_ISL_729320\|2020-1 | ···················· | ·A·················· | ···················· | ···················· | ···················· | ···A················ |
| Y | 2142 | hCoV-19/Canada/ON-S1905/2020\|EPI_ISL_729321\|2020-1 | ···················· | ·A·················· | ···················· | ···················· | ···················· | ···A················ |
| Y | 2143 | hCoV-19/Canada/ON-S1906/2020\|EPI_ISL_729322\|2020-1 | ···················· | ·A·················· | ···················· | ···················· | ···················· | ···A················ |
| Y | 2144 | hCoV-19/Canada/ON-S1908/2020\|EPI_ISL_729323\|2020-1 | ···················· | ·A·················· | ···················· | ···················· | ···················· | ···A················ |
| Y | 2145 | hCoV-19/Canada/ON-S1912/2020\|EPI_ISL_729324\|2020-1 | ···················· | ···················· | ···················· | ···················· | ···················· | ···················· |
| Y | 2146 | hCoV-19/Canada/ON-S1915/2020\|EPI_ISL_729325\|2020-1 | ···················· | ···················· | ···················· | ···················· | ···················· | ···················· |
| Y | 2147 | hCoV-19/Canada/ON-S1917/2020\|EPI_ISL_729326\|2020-1 | ···················· | ···················· | ···················· | ···················· | ···················· | ···················· |
| Y | 2148 | hCoV-19/Canada/ON-S1918/2020\|EPI_ISL_729327\|2020-1 | ···················· | ···················· | ···················· | ···················· | ···················· | ···················· |
| Y | 2149 | hCoV-19/Canada/ON-S1920/2020\|EPI_ISL_729328\|2020-1 | ···················· | ·A·················· | ···················· | ···················· | ···················· | ···A················ |
| Y | 2150 | hCoV-19/Canada/ON-S1921/2020\|EPI_ISL_729329\|2020-1 | ···················· | ·A·················· | ···················· | ···················· | ···················· | ···A················ |
| Y | 2151 | hCoV-19/Canada/ON-S1922/2020\|EPI_ISL_729330\|2020-1 | ···················· | ·A·················· | ···················· | ···················· | ···················· | ···A················ |
| Y | 2152 | hCoV-19/Canada/ON-S1925/2020\|EPI_ISL_729331\|2020-1 | ···················· | ···················· | ···················· | ···················· | ···················· | ···················· |
| Y | 2153 | hCoV-19/Canada/ON-S1930/2020\|EPI_ISL_729332\|2020-1 | ···················· | ···················· | ···················· | ············A······· | ···················· | ···················· |
| Y | 2154 | hCoV-19/Canada/ON-S1931/2020\|EPI_ISL_729333\|2020-1 | ···················· | ·A·················· | ···················· | ···················· | ···················· | ···A················ |
| Y | 2155 | hCoV-19/Canada/ON-S1932/2020\|EPI_ISL_729334\|2020-1 | ···················· | ···················· | ···················· | ···················· | ···················· | ···················· |
| Y | 2156 | hCoV-19/Canada/ON-S1934/2020\|EPI_ISL_729335\|2020-1 | ···················· | ···················· | ···················· | ···················· | ···················· | ···················· |
| Y | 2157 | hCoV-19/USA/CA-ALSR-5033/2020\|EPI_ISL_730304\|2020- | ···················· | ·A·················· | ···················· | ···················· | ···················· | ···················· |
| Y | 2158 | hCoV-19/USA/CA-ALSR-5035/2020\|EPI_ISL_730306\|2020- | ···················· | ·A·················· | ···················· | ···················· | ···················· | ···················· |
| Y | 2159 | hCoV-19/USA/CA-ALSR-5037/2020\|EPI_ISL_730307\|2020- | ···················· | ·A·················· | ···················· | ···················· | ···················· | ···················· |
| Y | 2160 | hCoV-19/USA/CA-ALSR-5039/2020\|EPI_ISL_730309\|2020- | ···················· | ·A·················· | ···················· | ···················· | ···················· | ···················· |
| Y | 2161 | hCoV-19/USA/CA-ALSR-5045/2020\|EPI_ISL_730312\|2020- | ···················· | ···················· | ···················· | ···················· | ···················· | ···················· |
| Y | 2162 | hCoV-19/USA/CA-ALSR-5046/2020\|EPI_ISL_730313\|2020- | ···················· | ···················· | ···················· | ···················· | ···················· | ···················· |
| Y | 2163 | hCoV-19/USA/CA-ALSR-5047/2020\|EPI_ISL_730314\|2020- | ···················· | ·A·················· | ···················· | ···················· | ···················· | ···················· |
| Y | 2164 | hCoV-19/USA/CA-ALSR-5048/2020\|EPI_ISL_730315\|2020- | ········T··········· | ·A·················· | ···················· | ···················· | ···················· | ···················· |
| Y | 2165 | hCoV-19/USA/CA-ALSR-5051/2020\|EPI_ISL_730318\|2020- | ···················· | ···················· | ···················· | ···················· | ···················· | ···················· |
| Y | 2166 | hCoV-19/USA/CA-ALSR-5052/2020\|EPI_ISL_730319\|2020- | ········T··········· | ·A·················· | ···················· | ···················· | ···················· | ···················· |
| Y | 2167 | hCoV-19/USA/CA-ALSR-5053/2020\|EPI_ISL_730320\|2020- | ···················· | ···················· | ···················· | ···················· | ···················· | ···················· |
| Y | 2168 | hCoV-19/USA/CA-ALSR-5055/2020\|EPI_ISL_730321\|2020- | ···················· | ·A·················· | ···················· | ···················· | ···················· | ···················· |
| Y | 2169 | hCoV-19/USA/CA-ALSR-5056/2020\|EPI_ISL_730322\|2020- | ···················· | ···················· | ···················· | ···················· | ···················· | ···················· |
| Y | 2170 | hCoV-19/USA/CA-ALSR-5057/2020\|EPI_ISL_730323\|2020- | ···················· | ···················· | ···················· | ···················· | ···················· | ···················· |
| Y | 2171 | hCoV-19/USA/CA-ALSR-5059/2020\|EPI_ISL_730324\|2020- | ···················· | ···················· | ···················· | ···················· | ···················· | ···················· |
| Y | 2172 | hCoV-19/USA/CA-ALSR-5060/2020\|EPI_ISL_730325\|2020- | ···················· | ·A·················· | ···················· | ···················· | ···················· | ···················· |
| Y | 2173 | hCoV-19/USA/CA-ALSR-5061/2020\|EPI_ISL_730326\|2020- | ···················· | ···················· | ···················· | ···················· | ···················· | ···················· |
| Y | 2174 | hCoV-19/USA/CA-ALSR-5064/2020\|EPI_ISL_730327\|2020- | ···················· | ·A·················· | ···················· | ···················· | ···················· | ···················· |
| Y | 2175 | hCoV-19/USA/CA-ALSR-5066/2020\|EPI_ISL_730328\|2020- | ···················· | ···················· | ···················· | ···················· | ···················· | ···················· |
| Y | 2176 | hCoV-19/USA/CA-ALSR-5069/2020\|EPI_ISL_730330\|2020- | ···················· | ·A·················· | ···················· | ···················· | ···················· | ···················· |
| Y | 2177 | hCoV-19/USA/CA-ALSR-5070/2020\|EPI_ISL_730331\|2020- | ···················· | ·A·················· | ···················· | ···················· | ···················· | ···················· |
| Y | 2178 | hCoV-19/USA/CA-ALSR-5071/2020\|EPI_ISL_730332\|2020- | ···················· | ·A·················· | ···················· | ···················· | ···················· | ···················· |
| Y | 2179 | hCoV-19/USA/CA-ALSR-5072/2020\|EPI_ISL_730333\|2020- | ···················· | ···················· | ···················· | ···················· | ···················· | ···················· |
| Y | 2180 | hCoV-19/USA/CA-ALSR-5073/2020\|EPI_ISL_730334\|2020- | ···················· | ·A·················· | ···················· | ···················· | ···················· | ···················· |
| Y | 2181 | hCoV-19/USA/CA-ALSR-5074/2020\|EPI_ISL_730335\|2020- | ···················· | ···················· | ···················· | ···················· | ···················· | ···················· |
| Y | 2182 | hCoV-19/USA/CA-ALSR-5077/2020\|EPI_ISL_730337\|2020- | ···················· | ···················· | ···················· | ···················· | ···················· | ···················· |
| Y | 2183 | hCoV-19/USA/CA-ALSR-5080/2020\|EPI_ISL_730338\|2020- | ···················· | ···················· | ···················· | ···················· | ···················· | ···················· |
| Y | 2184 | hCoV-19/USA/CA-ALSR-5082/2020\|EPI_ISL_730339\|2020- | ···················· | ···················· | ···················· | ···················· | ···················· | ···················· |

Y 2185 hCoV-19/USA/CA-ALSR-5083/2020|EPI_ISL_730340|2020- ···················· ·A·················· ····················· ···················· ···················· ····················
Y 2186 hCoV-19/USA/CA-ALSR-5084/2020|EPI_ISL_730341|2020- ···················· ···················· ····················· ···················· ···················· ····················
Y 2187 hCoV-19/USA/CA-ALSR-5087/2020|EPI_ISL_730342|2020- ···················· ·A·················· ····················· ···················· ···················· ····················
Y 2188 hCoV-19/USA/CA-ALSR-5088/2020|EPI_ISL_730343|2020- ···················· ···················· ····················· ···················· ···················· ····················
Y 2189 hCoV-19/USA/CA-ALSR-5089/2020|EPI_ISL_730344|2020- ···················· ···················· ····················· ···················· ···················· ····················
Y 2190 hCoV-19/USA/CA-ALSR-5090/2020|EPI_ISL_730345|2020- ···················· ···················· ····················· ···················· ···················· ····················
Y 2191 hCoV-19/USA/CA-ALSR-5093/2020|EPI_ISL_730347|2020- ···················· ·A·················· ····················· ···················· ···················· ····················
Y 2192 hCoV-19/USA/CA-ALSR-5096/2020|EPI_ISL_730348|2020- ···················· ···················· ····················· ···················· ···················· ····················
Y 2193 hCoV-19/USA/CA-ALSR-5097/2020|EPI_ISL_730349|2020- ···················· ·A·················· ····················· ···················· ···················· ····················
Y 2194 hCoV-19/USA/CA-ALSR-5100/2020|EPI_ISL_730351|2020- ···················· ·A·················· ····················· ···················· ···················· ····················
Y 2195 hCoV-19/USA/CA-ALSR-5104/2020|EPI_ISL_730353|2020- ···················· ···················· ····················· ···················· ···················· ····················
Y 2196 hCoV-19/USA/CA-ALSR-5107/2020|EPI_ISL_730354|2020- ···················· ·A·················· ····················· ···················· ···················· ····················
Y 2197 hCoV-19/USA/CA-ALSR-5108/2020|EPI_ISL_730355|2020- ···················· ·A·················· ····················· ···················· ···················· ····················
Y 2198 hCoV-19/USA/CA-ALSR-5109/2020|EPI_ISL_730356|2020- ···················· ·A·················· ····················· ···················· ···················· ····················
Y 2199 hCoV-19/USA/CA-ALSR-5110/2020|EPI_ISL_730357|2020- ···················· ·A·················· ····················· ···················· ···················· ····················
Y 2200 hCoV-19/USA/CA-ALSR-5111/2020|EPI_ISL_730358|2020- ···················· ·A·················· ····················· ···················· ···················· ····················
Y 2201 hCoV-19/USA/CA-ALSR-5112/2020|EPI_ISL_730359|2020- ···················· ···················· ····················· ···················· ···················· ····················
Y 2202 hCoV-19/USA/CA-ALSR-5115/2020|EPI_ISL_730361|2020- ···················· ·A·················· ····················· ···················· ···················· ····················
Y 2203 hCoV-19/USA/CA-ALSR-5116/2020|EPI_ISL_730362|2020- ···················· ·A·················· ····················· ···················· ···················· ····················
Y 2204 hCoV-19/USA/CA-ALSR-5118/2020|EPI_ISL_730363|2020- ···················· ·A·················· ····················· ···················· ···················· ····················
Y 2205 hCoV-19/USA/CA-ALSR-5121/2020|EPI_ISL_730364|2020- ···················· ···················· ····················· ···················· ···················· ····················
Y 2206 hCoV-19/USA/CA-ALSR-5122/2020|EPI_ISL_730365|2020- ···················· ·A·················· ····················· ···················· ···················· ····················
Y 2207 hCoV-19/USA/CA-ALSR-5123/2020|EPI_ISL_730366|2020- ···················· ·A·················· ····················· ···················· ···················· ····················
Y 2208 hCoV-19/USA/CA-ALSR-5127/2020|EPI_ISL_730368|2020- ···················· ·A·················· ····················· ···················· ···················· ····················
Y 2209 hCoV-19/USA/CA-ALSR-5128/2020|EPI_ISL_730369|2020- ···················· ···················· ····················· ···················· ···················· ····················
Y 2210 hCoV-19/USA/CA-ALSR-5129/2020|EPI_ISL_730370|2020- ···················· ···················· ····················· ···················· ···················· ····················
Y 2211 hCoV-19/Jordan/AM-ALSR-5132/2020|EPI_ISL_730372|20 ···················· ···················· ····················· ···················· ···················· ····················
Y 2212 hCoV-19/Jordan/AM-ALSR-5133/2020|EPI_ISL_730373|20 ···················· ···················· ····················· ···················· ···················· ····················
Y 2213 hCoV-19/Jordan/AM-ALSR-5134/2020|EPI_ISL_730374|20 ···················· ···················· ····················· ···················· ···················· ····················
Y 2214 hCoV-19/Jordan/AM-ALSR-5135/2020|EPI_ISL_730375|20 ···················· ···················· ····················· ···················· ···················· ····················
Y 2215 hCoV-19/Jordan/AM-ALSR-5136/2020|EPI_ISL_730376|20 ···················· ···················· ····················· ···················· ···················· ····················
Y 2216 hCoV-19/Jordan/AM-ALSR-5138/2020|EPI_ISL_730377|20 ···················· ···················· ····················· ···················· ···················· ····················
Y 2217 hCoV-19/Jordan/AM-ALSR-5139/2020|EPI_ISL_730378|20 ···················· ···················· ····················· ···················· ···················· ····················
Y 2218 hCoV-19/Jordan/AM-ALSR-5140/2020|EPI_ISL_730379|20 ···················· ···················· ····················· ···················· ···················· ····················
Y 2219 hCoV-19/Jordan/AM-ALSR-5141/2020|EPI_ISL_730380|20 ···················· ···················· ····················· ···················· ···················· ····················
Y 2220 hCoV-19/Jordan/AM-ALSR-5142/2020|EPI_ISL_730381|20 ···················· ···················· ····················· ···················· ···················· ····················
Y 2221 hCoV-19/Jordan/AM-ALSR-5143/2020|EPI_ISL_730382|20 ···················· ···················· ····················· ···················· ···················· ····················
Y 2222 hCoV-19/Jordan/AM-ALSR-5144/2020|EPI_ISL_730383|20 ···················· ·A·················· ····················· ···················· ···················· ··A·················
Y 2223 hCoV-19/Jordan/AM-ALSR-5145/2020|EPI_ISL_730384|20 ···················· ·A·················· ····················· ···················· ···················· ··A·················
Y 2224 hCoV-19/Jordan/AM-ALSR-5146/2020|EPI_ISL_730385|20 ···················· ·A·················· ····················· ···················· ···················· ··A·················
Y 2225 hCoV-19/Jordan/AM-ALSR-5147/2020|EPI_ISL_730386|20 ···················· ···················· ····················· ···················· ···················· ····················
Y 2226 hCoV-19/Jordan/AM-ALSR-5148/2020|EPI_ISL_730387|20 ···················· ···················· ····················· ···················· ···················· ····················
Y 2227 hCoV-19/Jordan/AM-ALSR-5149/2020|EPI_ISL_730388|20 ···················· ···················· ····················· ···················· ···················· ····················
Y 2228 hCoV-19/Jordan/AM-ALSR-5150/2020|EPI_ISL_730389|20 ···················· ···················· ····················· ···················· ···················· ····················
Y 2229 hCoV-19/England/ALDP-C3DAA6/2020|EPI_ISL_731634|20 ···················· ···················· ····················· ···················· ···················· ····················
Y 2230 hCoV-19/England/ALDP-C3C728/2020|EPI_ISL_731636|20 ···················· ···················· ····················· ···················· ···················· ····················
Y 2231 hCoV-19/England/ALDP-C3C0E1/2020|EPI_ISL_731638|20 ···················· ···················· ····················· ···················· ···················· ····················
Y 2232 hCoV-19/England/ALDP-C3A8FA/2020|EPI_ISL_731639|20 ···················· ···················· ····················· ···················· ···················· ····················
Y 2233 hCoV-19/England/ALDP-C3AAB8/2020|EPI_ISL_731640|20 ···················· ···················· ····················· ···················· ···················· ····················
Y 2234 hCoV-19/England/ALDP-C3AC1C/2020|EPI_ISL_731641|20 ···················· ···················· ····················· ···················· ···················· ····················
Y 2235 hCoV-19/England/ALDP-C3C6FE/2020|EPI_ISL_731642|20 ···················· ···················· ····················· ···················· ···················· ····················
Y 2236 hCoV-19/England/ALDP-C3CDEA/2020|EPI_ISL_731643|20 ···················· ···················· ····················· ···················· ···················· ····················
Y 2237 hCoV-19/England/ALDP-C3AFC8/2020|EPI_ISL_731644|20 ···················· ···················· ····················· ···················· ···················· ··A·················
Y 2238 hCoV-19/England/ALDP-C3CB68/2020|EPI_ISL_731645|20 ···················· ···················· ····················· ···················· ···················· ····················
Y 2239 hCoV-19/England/ALDP-C3D2EA/2020|EPI_ISL_731646|20 ···················· ···················· ····················· ···················· ···················· ····················
Y 2240 hCoV-19/England/ALDP-C3AFD7/2020|EPI_ISL_731649|20 ···················· ···················· ····················· ···················· ···················· ····················
Y 2241 hCoV-19/England/ALDP-C3BFF4/2020|EPI_ISL_731650|20 ···················· ···················· ····················· ···················· ···················· ····················
Y 2242 hCoV-19/England/ALDP-C3CE41/2020|EPI_ISL_731652|20 ···················· ···················· ····················· ···················· ···················· ····················
Y 2243 hCoV-19/England/ALDP-C3CDF9/2020|EPI_ISL_731653|20 ···················· ···················· ····················· ···················· ···················· ····················
Y 2244 hCoV-19/England/ALDP-C3BE8E/2020|EPI_ISL_731654|20 ···················· ···················· ····················· ···················· ···················· ····················
Y 2245 hCoV-19/England/ALDP-C3B11C/2020|EPI_ISL_731656|20 ···················· ·A·················· ····················· ···················· ···················· ··A·················
Y 2246 hCoV-19/England/ALDP-C3B8EA/2020|EPI_ISL_731659|20 ···················· ···················· ····················· ···················· ···················· ····················
Y 2247 hCoV-19/England/ALDP-C3C157/2020|EPI_ISL_731660|20 ···················· ···················· ····················· ···················· ······T············· ····················
Y 2248 hCoV-19/England/ALDP-C3D709/2020|EPI_ISL_731661|20 ···················· ···················· ····················· ···················· ···················· ····················
Y 2249 hCoV-19/England/ALDP-C3C6EF/2020|EPI_ISL_731662|20 ···················· ···················· ····················· ···················· ···················· ····················
Y 2250 hCoV-19/England/ALDP-C3D411/2020|EPI_ISL_731663|20 ···················· ···················· ····················· ···················· ···················· ····················
Y 2251 hCoV-19/England/ALDP-C3ADBF/2020|EPI_ISL_731667|20 ···················· ···················· ····················· ···················· ···················· ····················
Y 2252 hCoV-19/England/ALDP-C3C870/2020|EPI_ISL_731669|20 ···················· ···················· ···········T········· ···················· ···················· ····················
Y 2253 hCoV-19/England/ALDP-C3AFE6/2020|EPI_ISL_731671|20 ···················· ···················· ····················· ···················· ···················· ····················
Y 2254 hCoV-19/England/ALDP-C3BAC6/2020|EPI_ISL_731672|20 ···················· ···················· ····················· ···················· ···················· ····················
Y 2255 hCoV-19/England/ALDP-C3D5D2/2020|EPI_ISL_731675|20 ···················· ···················· ····················· ···················· ···················· ····················
Y 2256 hCoV-19/England/ALDP-C3D6A2/2020|EPI_ISL_731678|20 ···················· ···················· ····················· ···················· ···················· ····················
Y 2257 hCoV-19/England/ALDP-C3D6FD/2020|EPI_ISL_731679|20 ···················· ···················· ····················· ···················· ···················· ····················
Y 2258 hCoV-19/England/ALDP-C3D46C/2020|EPI_ISL_731681|20 ···················· ···················· ····················· ···················· ···················· ····················
Y 2259 hCoV-19/England/ALDP-C3BAB7/2020|EPI_ISL_731682|20 ···················· ···················· ····················· ···················· ···················· ····················
Y 2260 hCoV-19/England/ALDP-C3BC2A/2020|EPI_ISL_731683|20 ···················· ···················· ····················· ···················· ···················· ····················
Y 2261 hCoV-19/England/ALDP-C3A8CD/2020|EPI_ISL_731684|20 ···················· ···················· ····················· ···················· ···················· ····················
Y 2262 hCoV-19/England/ALDP-C3B0C4/2020|EPI_ISL_731686|20 ···················· ···················· ····················· ···················· ···················· ····················
Y 2263 hCoV-19/England/ALDP-C3B8BD/2020|EPI_ISL_731687|20 ···················· ···················· ····················· ···················· ···················· ····················
Y 2264 hCoV-19/England/ALDP-C3B2DD/2020|EPI_ISL_731691|20 ···················· ···················· ····················· ···················· ···················· ····················

| | | | | | | | | |
|---|---|---|---|---|---|---|---|---|
| Y | 2265 | hCoV-19/England/ALDP-C3ADDD/2020\|EPI_ISL_731692\|20 | .................... | .................... | .................... | .................... | .................... | .................... |
| Y | 2266 | hCoV-19/England/ALDP-C3CA5C/2020\|EPI_ISL_731694\|20 | .................... | .................... | .................... | .................... | .................... | .................... |
| Y | 2267 | hCoV-19/England/ALDP-C3DA79/2020\|EPI_ISL_731695\|20 | .................... | .................... | .................... | .................... | .................... | .................... |
| Y | 2268 | hCoV-19/England/ALDP-C3C89E/2020\|EPI_ISL_731696\|20 | .................... | .................... | .................... | .................... | .................... | .................... |
| Y | 2269 | hCoV-19/England/ALDP-C3B316/2020\|EPI_ISL_731697\|20 | .................... | .................... | .................... | .................... | .................... | .................... |
| Y | 2270 | hCoV-19/England/ALDP-C3C096/2020\|EPI_ISL_731698\|20 | .................... | .................... | .................... | .................... | .................... | .................... |
| Y | 2271 | hCoV-19/England/ALDP-C3BCA2/2020\|EPI_ISL_731699\|20 | .................... | .................... | .................... | .................... | .................... | .................... |
| Y | 2272 | hCoV-19/New Zealand/20CV0663/2020\|EPI_ISL_732965\|2 | .................... | .................... | .................... | .................... | .................... | .................... |
| Y | 2273 | hCoV-19/New Zealand/20CV0664/2020\|EPI_ISL_732966\|2 | .................... | .................... | .................... | .................... | .................... | .................... |
| Y | 2274 | hCoV-19/New Zealand/20CV0665/2020\|EPI_ISL_732967\|2 | .................... | .................... | .................... | .................... | .................... | .................... |
| Y | 2275 | hCoV-19/New Zealand/20CV0668/2020\|EPI_ISL_732968\|2 | .................... | .................... | .................... | .................... | .................... | .................... |
| Y | 2276 | hCoV-19/New Zealand/20CV0669/2020\|EPI_ISL_732969\|2 | .................... | .................... | .................... | .................... | .................... | .................... |
| Y | 2277 | hCoV-19/New Zealand/20CV0670/2020\|EPI_ISL_732970\|2 | .................... | .................... | .................... | .................... | .................... | .................... |
| Y | 2278 | hCoV-19/Russia/KR-RII-12754S/2020\|EPI_ISL_732975\|2 | .................... | .................... | .................... | .................... | .................... | .................... |
| Y | 2281 | hCoV-19/Russia/KR-RII-12221S/2020\|EPI_ISL_732980\|2 | .................... | .................... | .................... | .................... | .................... | .................... |
| Y | 2282 | hCoV-19/Russia/KR-RII-12211S/2020\|EPI_ISL_732982\|2 | .................... | .................... | .................... | .................... | .................... | .................... |
| Y | 2283 | hCoV-19/Russia/KR-RII-12210S/2020\|EPI_ISL_732983\|2 | .................... | .................... | .................... | .................... | .................... | .................... |
| Y | 2284 | hCoV-19/Russia/KR-RII-12198S/2020\|EPI_ISL_732985\|2 | .................... | .................... | .................... | .................... | N................... | .................... |
| Y | 2285 | hCoV-19/Russia/SPE-RII-10899S/2020\|EPI_ISL_732986\| | .................... | .................... | .................... | .................... | .................... | .................... |
| Y | 2286 | hCoV-19/Russia/SPE-RII-10016S/2020\|EPI_ISL_732987\| | .................... | .................... | .................... | .................... | .................... | .................... |
| Y | 2287 | hCoV-19/Russia/SPE-RII-9960S/2020\|EPI_ISL_732988\|2 | .................... | .................... | .................... | .................... | .................... | .................... |
| Y | 2288 | hCoV-19/Russia/SVE-RII-MH183S/2020\|EPI_ISL_732990\| | .................... | .................... | .................... | .................... | .................... | .................... |
| Y | 2289 | hCoV-19/Russia/SVE-RII-MH186S/2020\|EPI_ISL_732991\| | .................... | .................... | .................... | .................... | .................... | .................... |
| Y | 2290 | hCoV-19/Russia/SVE-RII-MH189S/2020\|EPI_ISL_732992\| | .................... | .................... | .................... | .................... | .................... | .................... |
| Y | 2291 | hCoV-19/Russia/SVE-RII-MH216S/2020\|EPI_ISL_732995\| | .................... | .................... | .................... | .................... | .................... | .................... |
| Y | 2292 | hCoV-19/Russia/SVE-RII-MH222S/2020\|EPI_ISL_732997\| | .................... | .................... | .................... | .................... | .................... | .................... |
| Y | 2293 | hCoV-19/Russia/SVE-RII-MH223S/2020\|EPI_ISL_732998\| | .................... | .................... | .................... | .................... | .................... | .................... |
| Y | 2294 | hCoV-19/Switzerland/SO-ETHZ-410836/2020\|EPI_ISL_73 | .................... | .................... | .................... | .................... | .................... | .................... |
| Y | 2295 | hCoV-19/Switzerland/SO-ETHZ-410841/2020\|EPI_ISL_73 | .................... | .................... | .................... | .................... | .................... | .................... |
| Y | 2297 | hCoV-19/Switzerland/SO-ETHZ-410846/2020\|EPI_ISL_73 | .................... | ......A............. | .................... | .................... | .................... | ...........M........ |
| Y | 2298 | hCoV-19/Switzerland/SO-ETHZ-410851/2020\|EPI_ISL_73 | .................... | .................... | .................... | .................... | .................... | .................... |
| Y | 2299 | hCoV-19/Switzerland/SO-ETHZ-410852/2020\|EPI_ISL_73 | .................... | .................... | .................... | .................... | .................... | .................... |
| Y | 2300 | hCoV-19/Switzerland/SO-ETHZ-410956/2020\|EPI_ISL_73 | .................... | .................... | .................... | .................... | .................... | .................... |
| Y | 2301 | hCoV-19/Switzerland/SO-ETHZ-410957/2020\|EPI_ISL_73 | .................... | .................... | .................... | .................... | .................... | .................... |
| Y | 2302 | hCoV-19/Switzerland/SO-ETHZ-410958/2020\|EPI_ISL_73 | .................... | .................... | .................... | .................... | .................... | .................... |
| Y | 2303 | hCoV-19/Switzerland/SG-ETHZ-410083/2020\|EPI_ISL_73 | .................... | .................... | .................... | .................... | .................... | .................... |
| Y | 2304 | hCoV-19/Switzerland/SG-ETHZ-410528/2020\|EPI_ISL_73 | .................... | .................... | .................... | .................... | .................... | .................... |
| Y | 2305 | hCoV-19/England/LIVE-DCC382/2020\|EPI_ISL_724501\|20 | .................... | .........T.......... | .................... | .................... | .................... | .................... |
| Y | 2306 | hCoV-19/England/LIVE-DCC434/2020\|EPI_ISL_724507\|20 | .................... | .................... | .................... | .................... | .................... | .................... |
| Y | 2307 | hCoV-19/England/LIVE-DCC4CB/2020\|EPI_ISL_724510\|20 | .................... | .................... | .................... | .................... | .................... | .................... |
| Y | 2308 | hCoV-19/England/LIVE-DCC531/2020\|EPI_ISL_724514\|20 | .................... | .................... | .................... | .................... | .................... | .................... |
| Y | 2309 | hCoV-19/England/LIVE-DCC55F/2020\|EPI_ISL_724516\|20 | .................... | .........T.......... | .................... | .................... | .................... | .................... |
| Y | 2310 | hCoV-19/England/LIVE-DCC5AA/2020\|EPI_ISL_724517\|20 | .................... | .................... | .................... | .................... | .................... | .................... |
| Y | 2311 | hCoV-19/England/LIVE-DCC5D7/2020\|EPI_ISL_724519\|20 | .................... | .................... | .................... | .................... | .................... | .................... |
| Y | 2312 | hCoV-19/England/LIVE-DCC62F/2020\|EPI_ISL_724522\|20 | .................... | .................... | .................... | .................... | .................... | .................... |
| Y | 2313 | hCoV-19/England/LOND-1264BF5/2020\|EPI_ISL_724545\|2 | .................... | .A.................. | .................... | .................... | .................... | ...A................ |
| Y | 2314 | hCoV-19/England/LOND-1264C01/2020\|EPI_ISL_724546\|2 | .................... | .................... | .................... | .................... | .................... | .................... |
| Y | 2315 | hCoV-19/England/LOND-1264C2F/2020\|EPI_ISL_724548\|2 | .................... | .................... | .................... | .................... | .................... | .................... |
| Y | 2316 | hCoV-19/England/LOND-1264C3E/2020\|EPI_ISL_724549\|2 | .................... | .................... | .................... | .................... | ......T............. | .................... |
| Y | 2317 | hCoV-19/England/LOND-1264C4D/2020\|EPI_ISL_724550\|2 | .................... | .................... | .................... | .................... | .................... | .................... |
| Y | 2318 | hCoV-19/England/LOND-1264C6B/2020\|EPI_ISL_724552\|2 | .................... | .A.................. | .................... | .................... | .................... | ...A................ |
| Y | 2319 | hCoV-19/England/LOND-1264CB6/2020\|EPI_ISL_724555\|2 | .................... | .................... | .................... | .................... | .................... | .................... |
| Y | 2320 | hCoV-19/England/LOND-1264CE3/2020\|EPI_ISL_724557\|2 | .................... | .................... | .................... | .................... | .................... | .................... |
| Y | 2321 | hCoV-19/England/LOND-1264D77/2020\|EPI_ISL_724565\|2 | .................... | .................... | .................... | .................... | .................... | .................... |
| Y | 2322 | hCoV-19/England/LOND-1264DC2/2020\|EPI_ISL_724568\|2 | .................... | .A.................. | .................... | .................... | .................... | .................... |
| Y | 2323 | hCoV-19/Wales/PHWC-492E8D/2020\|EPI_ISL_725800\|2020 | .................... | .................... | .................... | .................... | .................... | .................... |
| Y | 2324 | hCoV-19/Wales/PHWC-492E9C/2020\|EPI_ISL_725801\|2020 | .................... | .................... | .................... | .................... | .................... | .................... |
| Y | 2328 | hCoV-19/Wales/PHWC-492EE7/2020\|EPI_ISL_725805\|2020 | .................... | .................... | .................... | .................... | .................... | .................... |
| Y | 2329 | hCoV-19/Wales/PHWC-492EF6/2020\|EPI_ISL_725806\|2020 | .................... | .................... | .................... | .................... | .................... | .................... |
| Y | 2331 | hCoV-19/Wales/PHWC-492F4E/2020\|EPI_ISL_725811\|2020 | .................... | .................... | .................... | .................... | .................... | .................... |
| Y | 2332 | hCoV-19/Wales/PHWC-492F5D/2020\|EPI_ISL_725812\|2020 | .................... | .................... | .................... | .................... | .................... | .................... |
| Y | 2333 | hCoV-19/Wales/PHWC-492F6C/2020\|EPI_ISL_725813\|2020 | .................... | .................... | .................... | .................... | .................... | .................... |
| Y | 2334 | hCoV-19/Wales/PHWC-492F7B/2020\|EPI_ISL_725814\|2020 | .................... | .................... | .................... | .................... | .................... | .................... |
| Y | 2335 | hCoV-19/Wales/PHWC-49303B/2020\|EPI_ISL_725826\|2020 | .................... | .................... | .................... | .................... | NNN................. | .................... | 1 |
| Y | 2336 | hCoV-19/Wales/PHWC-49304A/2020\|EPI_ISL_725827\|2020 | .................... | .................... | .................... | .................... | .................... | .................... |
| Y | 2337 | hCoV-19/Wales/PHWC-493059/2020\|EPI_ISL_725828\|2020 | .................... | .................... | .................... | .................... | .................... | .................... |
| Y | 2339 | hCoV-19/Wales/PHWC-493077/2020\|EPI_ISL_725830\|2020 | .................... | .................... | .................... | .................... | .................... | .................... |
| Y | 2340 | hCoV-19/Wales/PHWC-493086/2020\|EPI_ISL_725831\|2020 | .................... | .................... | .................... | .................... | .................... | .................... |
| Y | 2341 | hCoV-19/Wales/PHWC-493095/2020\|EPI_ISL_725832\|2020 | .................... | .................... | .................... | .................... | .................... | .................... |
| Y | 2342 | hCoV-19/Wales/PHWC-4930E0/2020\|EPI_ISL_725837\|2020 | .................... | .................... | .................... | .................... | .................... | .................... |
| Y | 2343 | hCoV-19/Wales/PHWC-4930FF/2020\|EPI_ISL_725838\|2020 | .................... | .................... | .................... | .................... | .................... | .................... |
| Y | 2344 | hCoV-19/Wales/PHWC-49310B/2020\|EPI_ISL_725839\|2020 | .................... | .................... | .................... | .................... | .................... | .................... |
| Y | 2345 | hCoV-19/Wales/PHWC-49311A/2020\|EPI_ISL_725840\|2020 | .................... | .................... | .................... | .................... | .................... | .................... |
| Y | 2346 | hCoV-19/Wales/PHWC-493129/2020\|EPI_ISL_725841\|2020 | .................... | .................... | .................... | .................... | .................... | .................... |
| Y | 2347 | hCoV-19/Wales/PHWC-493138/2020\|EPI_ISL_725842\|2020 | .................... | .................... | .................... | .................... | .................... | .................... |
| Y | 2348 | hCoV-19/Wales/PHWC-493156/2020\|EPI_ISL_725844\|2020 | .................... | .................... | .................... | .................... | .................... | .................... |
| Y | 2349 | hCoV-19/Wales/PHWC-493165/2020\|EPI_ISL_725845\|2020 | .................... | .................... | .................... | .................... | .................... | .................... |
| Y | 2350 | hCoV-19/Wales/PHWC-493174/2020\|EPI_ISL_725846\|2020 | .................... | .................... | .................... | .................... | .................... | .................... |
| Y | 2351 | hCoV-19/Wales/PHWC-493183/2020\|EPI_ISL_725847\|2020 | .................... | .................... | .................... | .................... | .................... | .................... |
| Y | 2353 | hCoV-19/Wales/PHWC-4931A1/2020\|EPI_ISL_725849\|2020 | .................... | .................... | .................... | .................... | .................... | .................... |

| | | | | | | | | |
|---|---|---|---|---|---|---|---|---|
| Y | 2354 | hCoV-19/Wales/PHWC-4931B0/2020\|EPI_ISL_725850\|2020 | ···················· | ···················· | ···················· | ···················· | ···················· | ···················· |
| Y | 2355 | hCoV-19/Wales/PHWC-4931CF/2020\|EPI_ISL_725851\|2020 | ···················· | ···················· | ···················· | ···················· | ···················· | ···················· |
| Y | 2356 | hCoV-19/Wales/PHWC-4931DE/2020\|EPI_ISL_725852\|2020 | ···················· | ···················· | ···················· | ···················· | ···················· | ···················· |
| Y | 2357 | hCoV-19/Wales/PHWC-4931ED/2020\|EPI_ISL_725853\|2020 | ···················· | ···················· | ···················· | ···················· | ···················· | ···················· |
| Y | 2358 | hCoV-19/Wales/PHWC-4931FC/2020\|EPI_ISL_725854\|2020 | ···················· | ···················· | ···················· | ···················· | ···················· | ···················· |
| Y | 2359 | hCoV-19/Wales/PHWC-493208/2020\|EPI_ISL_725855\|2020 | ···················· | ···················· | ···················· | ···················· | ···················· | ···················· |
| Y | 2362 | hCoV-19/Wales/PHWC-493280/2020\|EPI_ISL_725861\|2020 | ···················· | ···················· | ···················· | ···················· | ···················· | ···················· |
| Y | 2364 | hCoV-19/Wales/PHWC-4932BD/2020\|EPI_ISL_725864\|2020 | ···················· | ···················· | ···················· | ···················· | ···················· | ···················· |
| Y | 2366 | hCoV-19/Wales/PHWC-4932EA/2020\|EPI_ISL_725866\|2020 | ···················· | ···················· | ···················· | ···················· | ···················· | ···················· |
| Y | 2367 | hCoV-19/Wales/PHWC-4932F9/2020\|EPI_ISL_725867\|2020 | ···················· | ···················· | ···················· | ···················· | ···················· | ···················· |
| Y | 2369 | hCoV-19/Wales/PHWC-493314/2020\|EPI_ISL_725869\|2020 | ···················· | ···················· | ···················· | ···················· | ···················· | ···················· |
| Y | 2370 | hCoV-19/Wales/PHWC-493332/2020\|EPI_ISL_725870\|2020 | ···················· | ···················· | ···················· | ···················· | ···················· | ···················· |
| Y | 2372 | hCoV-19/Wales/PHWC-49336F/2020\|EPI_ISL_725873\|2020 | ···················· | ···················· | ···················· | ···················· | ···················· | ···················· |
| Y | 2374 | hCoV-19/Wales/PHWC-49338D/2020\|EPI_ISL_725875\|2020 | ···················· | ···················· | ···················· | ···················· | ···················· | ···················· |
| Y | 2375 | hCoV-19/Wales/PHWC-49339C/2020\|EPI_ISL_725876\|2020 | ···················· | ···················· | ···················· | ···················· | ···················· | ···················· |
| Y | 2377 | hCoV-19/Wales/PHWC-4933BA/2020\|EPI_ISL_725878\|2020 | ···················· | ···················· | ···················· | ···················· | ···················· | ···················· |
| Y | 2379 | hCoV-19/Wales/PHWC-4933F6/2020\|EPI_ISL_725882\|2020 | ···················· | ···················· | ···················· | ···················· | ···················· | ···················· |
| Y | 2380 | hCoV-19/Wales/PHWC-493402/2020\|EPI_ISL_725883\|2020 | ···················· | ···················· | ···················· | ···················· | ···················· | ···················· |
| Y | 2381 | hCoV-19/Wales/PHWC-493411/2020\|EPI_ISL_725884\|2020 | ···················· | ···················· | ···················· | ···················· | ···················· | ···················· |
| Y | 2383 | hCoV-19/Wales/PHWC-49343F/2020\|EPI_ISL_725886\|2020 | ···················· | ···················· | ···················· | ···················· | ···················· | ···················· |
| Y | 2384 | hCoV-19/Wales/PHWC-49344E/2020\|EPI_ISL_725887\|2020 | ···················· | ···················· | ···················· | ···················· | ···················· | ···················· |
| Y | 2385 | hCoV-19/Wales/PHWC-49345D/2020\|EPI_ISL_725888\|2020 | ···················· | ···················· | ···················· | ···················· | ···················· | ···················· |
| Y | 2386 | hCoV-19/Wales/PHWC-49346C/2020\|EPI_ISL_725889\|2020 | ···················· | ···················· | ···················· | ···················· | ···················· | ···················· |
| Y | 2388 | hCoV-19/Wales/PHWC-4934E4/2020\|EPI_ISL_725897\|2020 | ···················· | ···················· | ···················· | ···················· | ···················· | ···················· |
| Y | 2389 | hCoV-19/Wales/PHWC-4934F3/2020\|EPI_ISL_725898\|2020 | ···················· | ···················· | ···················· | ···················· | ···················· | ···················· |
| Y | 2390 | hCoV-19/Wales/PHWC-49350F/2020\|EPI_ISL_725899\|2020 | ···················· | ···················· | ···················· | ···················· | ···················· | ···················· |
| Y | 2392 | hCoV-19/USA/WI-UW-2305/2020\|EPI_ISL_728071\|2020-11 | ···················· | ···················· | ···················· | ···················· | ···················· | ···················· |
| Y | 2401 | hCoV-19/USA/WI-UW-2314/2020\|EPI_ISL_728080\|2020-11 | ···················· | ···················· | ···················· | ···················· | ···················· | ···················· |
| Y | 2403 | hCoV-19/USA/WI-UW-2316/2020\|EPI_ISL_728082\|2020-11 | ···················· | ·A·················· | ···················· | ···················· | ···················· | ···················· |
| Y | 2406 | hCoV-19/USA/WI-UW-2320/2020\|EPI_ISL_728086\|2020-11 | ···················· | ···················· | ···················· | ···················· | ···················· | ···················· |
| Y | 2407 | hCoV-19/USA/WI-UW-2321/2020\|EPI_ISL_728087\|2020-11 | ···················· | ···················· | ···················· | ···················· | ···················· | ···················· |
| Y | 2408 | hCoV-19/USA/WI-UW-2322/2020\|EPI_ISL_728088\|2020-10 | ···················· | ···················· | ···················· | ···················· | ···················· | ···················· |
| Y | 2409 | hCoV-19/USA/WI-UW-2323/2020\|EPI_ISL_728089\|2020-12 | ···················· | ···················· | ···················· | ···················· | ···················· | ···················· |
| Y | 2410 | hCoV-19/Jordan/AM-ALSR-5151/2020\|EPI_ISL_730390\|20 | ···················· | ···················· | ···················· | ···················· | ···················· | ···················· |
| Y | 2411 | hCoV-19/Jordan/AM-ALSR-5152/2020\|EPI_ISL_730391\|20 | ···················· | ···················· | ···················· | ···················· | ···················· | ···················· |
| Y | 2412 | hCoV-19/Jordan/AM-ALSR-5153/2020\|EPI_ISL_730392\|20 | ···················· | ·A·················· | ···················· | ···················· | ···················· | ··A················· |
| Y | 2413 | hCoV-19/Jordan/AM-ALSR-5154/2020\|EPI_ISL_730393\|20 | ···················· | ···················· | ···················· | ···················· | ···················· | ···················· |
| Y | 2414 | hCoV-19/Jordan/AM-ALSR-5155/2020\|EPI_ISL_730394\|20 | ···················· | ···················· | ···················· | ···················· | ···················· | ···················· |
| Y | 2415 | hCoV-19/Jordan/AM-ALSR-5156/2020\|EPI_ISL_730395\|20 | ···················· | ···················· | ···················· | ···················· | ···················· | ···················· |
| Y | 2416 | hCoV-19/Jordan/AM-ALSR-5157/2020\|EPI_ISL_730396\|20 | ···················· | ···················· | ···················· | ···················· | ···················· | ···················· |
| Y | 2417 | hCoV-19/Jordan/AM-ALSR-5158/2020\|EPI_ISL_730397\|20 | ···················· | ···················· | ···················· | ···················· | ···················· | ···················· |
| Y | 2418 | hCoV-19/Jordan/AM-ALSR-5159/2020\|EPI_ISL_730398\|20 | ···················· | ·A·················· | ···················· | ···················· | ···················· | ··A················· |
| Y | 2419 | hCoV-19/Jordan/AM-ALSR-5160/2020\|EPI_ISL_730399\|20 | ···················· | ···················· | ···················· | ···················· | ···················· | ···················· |
| Y | 2420 | hCoV-19/Belgium/rega-10191306/2020\|EPI_ISL_735209\| | ···················· | ···················· | ···················· | ···················· | ···················· | ···················· |
| Y | 2421 | hCoV-19/Belgium/rega-10201104/2020\|EPI_ISL_735210\| | ···················· | ···················· | ···················· | ···················· | ···················· | ···················· |
| Y | 2422 | hCoV-19/Belgium/rega-10201105/2020\|EPI_ISL_735211\| | ···················· | ···················· | ···················· | ···················· | ···················· | ···················· |
| Y | 2423 | hCoV-19/Belgium/rega-10201106/2020\|EPI_ISL_735212\| | ···················· | ···················· | ···················· | ···················· | ···················· | ···················· |
| Y | 2424 | hCoV-19/Belgium/rega-10201108/2020\|EPI_ISL_735213\| | ···················· | ···················· | ···················· | ···················· | ···················· | ···················· |
| Y | 2425 | hCoV-19/Belgium/rega-10201110/2020\|EPI_ISL_735214\| | ···················· | ···················· | ···················· | ···················· | ···················· | ···················· |
| Y | 2426 | hCoV-19/Belgium/rega-11021204/2020\|EPI_ISL_735215\| | ···················· | ···················· | ···················· | ···················· | ···················· | ···················· |
| Y | 2427 | hCoV-19/Belgium/rega-11021206/2020\|EPI_ISL_735216\| | ···················· | ···················· | ···················· | ···················· | ···················· | ···················· |
| Y | 2428 | hCoV-19/Belgium/rega-11031205/2020\|EPI_ISL_735217\| | ···················· | ···················· | ···················· | ···················· | ···················· | ···················· |
| Y | 2429 | hCoV-19/Belgium/rega-11031207/2020\|EPI_ISL_735218\| | ···················· | ···················· | ···················· | ···················· | ···················· | ···················· |
| Y | 2430 | hCoV-19/Belgium/rega-11031208/2020\|EPI_ISL_735219\| | ···················· | ···················· | ···················· | ···················· | ···················· | ···················· |
| Y | 2431 | hCoV-19/Belgium/rega-11031209/2020\|EPI_ISL_735220\| | ···················· | ···················· | ···················· | ···················· | ···················· | ···················· |
| Y | 2432 | hCoV-19/Belgium/rega-11271168/2020\|EPI_ISL_735221\| | ···················· | ···················· | ···················· | ···················· | ···················· | ···················· |
| Y | 2433 | hCoV-19/Belgium/rega-11271169/2020\|EPI_ISL_735222\| | ···················· | ···················· | ···················· | ···················· | ···················· | ···················· |
| Y | 2434 | hCoV-19/Belgium/rega-11271170/2020\|EPI_ISL_735223\| | ···················· | ···················· | ···················· | ···················· | ···················· | ···················· |
| Y | 2435 | hCoV-19/Belgium/rega-11271171/2020\|EPI_ISL_735224\| | ···················· | ···················· | ···················· | ···················· | ···················· | ···················· |
| Y | 2436 | hCoV-19/Belgium/rega-11271172/2020\|EPI_ISL_735225\| | ···················· | ···················· | ···················· | ···················· | ···················· | ···················· |
| Y | 2437 | hCoV-19/Belgium/rega-11271173/2020\|EPI_ISL_735226\| | ···················· | ···················· | ···················· | ···················· | ···················· | ···················· |
| Y | 2438 | hCoV-19/Belgium/rega-11271174/2020\|EPI_ISL_735227\| | ···················· | ···················· | ···················· | ···················· | ···················· | ···················· |
| Y | 2439 | hCoV-19/Belgium/rega-11271175/2020\|EPI_ISL_735228\| | ···················· | ···················· | ···················· | ···················· | ···················· | ···················· |
| Y | 2440 | hCoV-19/Belgium/rega-11271176/2020\|EPI_ISL_735229\| | ···················· | ···················· | ···················· | ···················· | ···················· | ···················· |
| Y | 2441 | hCoV-19/Belgium/rega-11271177/2020\|EPI_ISL_735230\| | ···················· | ···················· | ···················· | ···················· | ···················· | ···················· |
| Y | 2442 | hCoV-19/Belgium/rega-11271178/2020\|EPI_ISL_735231\| | ···················· | ···················· | ···················· | ···················· | ···················· | ···················· |
| Y | 2443 | hCoV-19/Belgium/rega-11271179/2020\|EPI_ISL_735232\| | ···················· | ···················· | ···················· | ···················· | ···················· | ···················· |
| Y | 2444 | hCoV-19/Belgium/rega-11271180/2020\|EPI_ISL_735233\| | ···················· | ···················· | ···················· | ···················· | ···················· | ···················· |
| Y | 2445 | hCoV-19/Belgium/rega-11271181/2020\|EPI_ISL_735234\| | ···················· | ···················· | ···················· | ···················· | ···················· | ···················· |
| Y | 2446 | hCoV-19/Belgium/rega-11271182/2020\|EPI_ISL_735235\| | ···················· | ···················· | ···················· | ···················· | ···················· | ···················· |
| Y | 2447 | hCoV-19/Belgium/rega-11301199/2020\|EPI_ISL_735236\| | ···················· | ···················· | ···················· | ···················· | ···················· | ···················· |
| Y | 2448 | hCoV-19/Belgium/rega-11301201/2020\|EPI_ISL_735237\| | ···················· | ···················· | ···················· | ···················· | ···················· | ···················· |
| Y | 2449 | hCoV-19/Belgium/rega-11301203/2020\|EPI_ISL_735238\| | ···················· | ···················· | ···················· | ···················· | ···················· | ···················· |
| Y | 2450 | hCoV-19/Belgium/rega-12011200/2020\|EPI_ISL_735239\| | ···················· | ···················· | ···················· | ···················· | ···················· | ···················· |
| Y | 2451 | hCoV-19/England/ALDP-C4862C/2020\|EPI_ISL_736500\|20 | ···················· | ···················· | ···················· | ···················· | ···················· | ···················· |
| Y | 2452 | hCoV-19/England/MILK-C50393/2020\|EPI_ISL_736502\|20 | ···················· | ···················· | ···················· | ···················· | ···················· | ···················· |
| Y | 2453 | hCoV-19/England/MILK-C508C1/2020\|EPI_ISL_736504\|20 | ···················· | ···················· | ···················· | ···················· | ···················· | ···················· |
| Y | 2454 | hCoV-19/England/ALDP-C48CDF/2020\|EPI_ISL_736506\|20 | ···················· | ···················· | ···················· | ···················· | ···················· | ···················· |
| Y | 2455 | hCoV-19/England/ALDP-C48BF0/2020\|EPI_ISL_736510\|20 | ···················· | ···················· | ···················· | ···················· | ···················· | ···················· |
| Y | 2456 | hCoV-19/England/ALDP-C48B78/2020\|EPI_ISL_736511\|20 | ···················· | ···················· | ···················· | ···················· | ···················· | ···················· |

```
Y 2457 hCoV-19/England/ALDP-C48853/2020|EPI_ISL_736513|20 ···················· ···················· ····················· ···················· ···················· ····················
Y 2458 hCoV-19/England/ALDP-C48686/2020|EPI_ISL_736517|20 ···················· ···················· ····················· ···················· ······T············· ····················
Y 2459 hCoV-19/England/ALDP-C48862/2020|EPI_ISL_736518|20 ···················· ···················· ····················· ···················· ···················· ····················
Y 2460 hCoV-19/England/ALDP-C48747/2020|EPI_ISL_736520|20 ···················· ···················· ····················· ···················· ···················· ····················
Y 2461 hCoV-19/England/ALDP-C487CF/2020|EPI_ISL_736522|20 ···················· ···················· ····················· ···················· ···················· ····················
Y 2462 hCoV-19/England/MILK-C50366/2020|EPI_ISL_736523|20 ···················· ···················· ····················· ···················· ···················· ····················
Y 2463 hCoV-19/England/ALDP-C48598/2020|EPI_ISL_736526|20 ···················· ·A·················· ····················· ···················· ···················· ··A·················
Y 2464 hCoV-19/England/MILK-C50454/2020|EPI_ISL_736527|20 ···················· ···················· ····················· ···················· ···················· ····················
Y 2465 hCoV-19/England/ALDP-C48B2D/2020|EPI_ISL_736540|20 ···················· ···················· ····················· ···················· ···················· ····················
Y 2466 hCoV-19/England/ALDP-C48677/2020|EPI_ISL_736541|20 ···················· ···················· ····················· ···················· ···················· ····················
Y 2467 hCoV-19/England/MILK-C504DC/2020|EPI_ISL_736542|20 ···················· ·A·················· ····················· ···················· ···················· ··A·················
Y 2468 hCoV-19/England/ALDP-C48D45/2020|EPI_ISL_736543|20 ···················· ···················· ····················· ···················· ···················· ····················
Y 2469 hCoV-19/England/ALDP-C485A7/2020|EPI_ISL_736545|20 ···················· ·A·················· ····················· ···················· ···················· ··A·················
Y 2470 hCoV-19/England/ALDP-C48A5D/2020|EPI_ISL_736548|20 ···················· ···················· ····················· ···················· ···················· ····················
Y 2471 hCoV-19/England/MILK-C50603/2020|EPI_ISL_736550|20 ···················· ···················· ····················· ···················· ···················· ····················
Y 2472 hCoV-19/England/MILK-C50445/2020|EPI_ISL_736551|20 ···················· ·A·················· ····················· ···················· ···················· ····················
Y 2473 hCoV-19/England/MILK-C503C0/2020|EPI_ISL_736552|20 ··················T· ···················· ····················· ···················· ···················· ···················· 1
Y 2474 hCoV-19/England/ALDP-C488F9/2020|EPI_ISL_736557|20 ···················· ···················· ····················· ···················· ······T············· ····················
Y 2475 hCoV-19/England/ALDP-C48A4E/2020|EPI_ISL_736559|20 ···················· ···················· ····················· ···················· ···················· ····················
Y 2476 hCoV-19/England/ALDP-C48B5A/2020|EPI_ISL_736560|20 ···················· ···················· ····················· ···················· ···················· ····················
Y 2477 hCoV-19/England/ALDP-C48DBE/2020|EPI_ISL_736561|20 ···················· ···················· ····················· ···················· ···················· ····················
Y 2478 hCoV-19/England/ALDP-C48440/2020|EPI_ISL_736562|20 ···················· ···················· ····················· ···················· ···················· ····················
Y 2479 hCoV-19/England/MILK-C503DF/2020|EPI_ISL_736565|20 ···················· ···················· ····················· ···················· ···················· ····················
Y 2480 hCoV-19/England/QEUH-C4BCFA/2020|EPI_ISL_736567|20 ···················· ···················· ····················· ···················· ···················· ····················
Y 2481 hCoV-19/England/ALDP-C48941/2020|EPI_ISL_736569|20 ···················· ···················· ····················· ···················· ···················· ····················
Y 2482 hCoV-19/Switzerland/SO-ETHZ-410744/2020|EPI_ISL_73 ···················· ···················· ····················· ···················· ···················· ····················
Y 2483 hCoV-19/Switzerland/SO-ETHZ-410746/2020|EPI_ISL_73 ···················· ···················· ····················· ···················· ···················· ····················
Y 2484 hCoV-19/Switzerland/SO-ETHZ-410828/2020|EPI_ISL_73 ···················· ···················· ····················· ···················· ···················· ····················
Y 2485 hCoV-19/Switzerland/SO-ETHZ-410829/2020|EPI_ISL_73 ···················· ···················· ····················· ···················· ···················· ····················
Y 2487 hCoV-19/Switzerland/SG-ETHZ-410901/2020|EPI_ISL_73 ···················· ···················· ····················· ···················· ···················· ····················
Y 2488 hCoV-19/Switzerland/TG-ETHZ-410084/2020|EPI_ISL_73 ···················· ···················· ····················· ···················· ···················· ····················
Y 2489 hCoV-19/Switzerland/TG-ETHZ-410086/2020|EPI_ISL_73 ···················· ···················· ····················· ···················· ···················· ····················
Y 2490 hCoV-19/Switzerland/TG-ETHZ-410087/2020|EPI_ISL_73 ···················· ···················· ····················· ···················· ···················· ····················
Y 2491 hCoV-19/Switzerland/TI-ETHZ-410125/2020|EPI_ISL_73 ···················· ···················· ····················· ···················· ···················· ····················
Y 2492 hCoV-19/Switzerland/TI-ETHZ-410133/2020|EPI_ISL_73 ···················· ···················· ····················· ···················· ···················· ····················
Y 2493 hCoV-19/Switzerland/VS-ETHZ-320189/2020|EPI_ISL_73 ···················· ···················· ····················· ···················· ···················· ····················
Y 2494 hCoV-19/Switzerland/VS-ETHZ-320190/2020|EPI_ISL_73 ···················· ···················· ····················· ···················· ···················· ····················
Y 2495 hCoV-19/Switzerland/VS-ETHZ-320192/2020|EPI_ISL_73 ···················· ···················· ····················· ···················· ···················· ····················
Y 2496 hCoV-19/Switzerland/VS-ETHZ-320194/2020|EPI_ISL_73 ···················· ···················· ····················· ···················· ···················· ····················
Y 2497 hCoV-19/Switzerland/VS-ETHZ-320196/2020|EPI_ISL_73 ···················· ···················· ····················· ···················· ···················· ····················
Y 2500 hCoV-19/Switzerland/VS-ETHZ-320205/2020|EPI_ISL_73 ···················· ···················· ····················· ···················· ···················· ····················
Y 2502 hCoV-19/Switzerland/VS-ETHZ-320207/2020|EPI_ISL_73 ···················· ···················· ····················· ···················· ···················· ····················
Y 2503 hCoV-19/Switzerland/VS-ETHZ-320401/2020|EPI_ISL_73 ···················· ···················· ····················· ···················· ···················· ····················
Y 2504 hCoV-19/Switzerland/VS-ETHZ-320408/2020|EPI_ISL_73 ···················· ···················· ····················· ···················· ···················· ····················
Y 2505 hCoV-19/Switzerland/VS-ETHZ-320410/2020|EPI_ISL_73 ···················· ···················· ····················· ···················· ···················· ····················
Y 2506 hCoV-19/Switzerland/VS-ETHZ-320412/2020|EPI_ISL_73 ···················· ···················· ····················· ···················· ···················· ····················
Y 2508 hCoV-19/Switzerland/VS-ETHZ-320586/2020|EPI_ISL_73 ···················· ···················· ····················· ···················· ···················· ····················
Y 2509 hCoV-19/Switzerland/VS-ETHZ-320592/2020|EPI_ISL_73 ···················· ···················· ····················· ···················· ···················· ····················
Y 2510 hCoV-19/Switzerland/VS-ETHZ-360051/2020|EPI_ISL_73 ···················· ·A·················· ····················· ···················· ···················· ····················
Y 2511 hCoV-19/Switzerland/VS-ETHZ-410031/2020|EPI_ISL_73 ···················· ···················· ····················· ···················· ···················· ····················
Y 2512 hCoV-19/Switzerland/VS-ETHZ-410052/2020|EPI_ISL_73 ···················· ···················· ····················· ···················· ···················· ····················
Y 2513 hCoV-19/Switzerland/VS-ETHZ-410062/2020|EPI_ISL_73 ···················· ···················· ····················· ···················· ···················· ····················
Y 2514 hCoV-19/Switzerland/VS-ETHZ-410068/2020|EPI_ISL_73 ···················· ···················· ····················· ···················· ···················· ····················
Y 2515 hCoV-19/Switzerland/VS-ETHZ-410095/2020|EPI_ISL_73 ···················· ···················· ····················· ···················· ···················· ····················
Y 2516 hCoV-19/Switzerland/VS-ETHZ-410148/2020|EPI_ISL_73 ···················· ···················· ····················· ···················· ···················· ····················
Y 2517 hCoV-19/Switzerland/VS-ETHZ-410149/2020|EPI_ISL_73 ···················· ···················· ····················· ···················· ···················· ····················
Y 2518 hCoV-19/Switzerland/VS-ETHZ-410154/2020|EPI_ISL_73 ···················· ···················· ····················· ···················· ···················· ····················
Y 2519 hCoV-19/Switzerland/VS-ETHZ-410248/2020|EPI_ISL_73 ···················· ···················· ····················· ···················· ···················· ····················
Y 2520 hCoV-19/Switzerland/VS-ETHZ-410350/2020|EPI_ISL_73 ···················· ···················· ····················· ···················· ···················· ····················
Y 2521 hCoV-19/Switzerland/VS-ETHZ-410378/2020|EPI_ISL_73 ···················· ···················· ····················· ···················· ···················· ····················
Y 2522 hCoV-19/Switzerland/VS-ETHZ-410386/2020|EPI_ISL_73 ···················· ···················· ····················· ···················· ···················· ····················
Y 2523 hCoV-19/Switzerland/VS-ETHZ-410411/2020|EPI_ISL_73 ···················· ···················· ····················· ···················· ···················· ····················
Y 2524 hCoV-19/Switzerland/VS-ETHZ-410436/2020|EPI_ISL_73 ···················· ···················· ····················· ···················· ···················· ····················
Y 2525 hCoV-19/Switzerland/VS-ETHZ-410494/2020|EPI_ISL_73 ···················· ···················· ····················· ···················· ···················· ····················
Y 2526 hCoV-19/Switzerland/VS-ETHZ-410544/2020|EPI_ISL_73 ···················· ···················· ····················· ···················· ···················· ····················
Y 2527 hCoV-19/Switzerland/VS-ETHZ-410559/2020|EPI_ISL_73 ···················· ···················· ····················· ···················· ···················· ····················
Y 2528 hCoV-19/Switzerland/VS-ETHZ-410573/2020|EPI_ISL_73 ···················· ···················· ····················· ···················· ············K······· ····················
Y 2529 hCoV-19/Switzerland/VS-ETHZ-410604/2020|EPI_ISL_73 ···················· ···················· ····················· ···················· ···················· ····················
Y 2530 hCoV-19/Switzerland/VS-ETHZ-410625/2020|EPI_ISL_73 ···················· ···················· ····················· ···················· ···················· ····················
Y 2531 hCoV-19/Switzerland/VS-ETHZ-410872/2020|EPI_ISL_73 ···················· ···················· ····················· ···················· ···················· ····················
Y 2532 hCoV-19/Switzerland/VS-ETHZ-410899/2020|EPI_ISL_73 ···················· ···················· ····················· ···················· ···················· ····················
Y 2533 hCoV-19/Switzerland/VS-ETHZ-410903/2020|EPI_ISL_73 ···················· ···················· ····················· ···················· ···················· ····················
Y 2534 hCoV-19/Switzerland/VS-ETHZ-410921/2020|EPI_ISL_73 ···················· ···················· ····················· ···················· ···················· ····················
Y 2535 hCoV-19/Switzerland/VS-ETHZ-410990/2020|EPI_ISL_73 ···················· ···················· ····················· ···················· ···················· ····················
Y 2536 hCoV-19/Switzerland/VS-ETHZ-411013/2020|EPI_ISL_73 ···················· ···················· ····················· ···················· ···················· ····················
Y 2537 hCoV-19/Switzerland/VS-ETHZ-411027/2020|EPI_ISL_73 ···················· ···················· ····················· ···················· ···················· ····················
Y 2538 hCoV-19/Switzerland/VS-ETHZ-411038/2020|EPI_ISL_73 ···················· ···················· ····················· ···················· ···················· ····················
Y 2539 hCoV-19/Switzerland/VS-ETHZ-411040/2020|EPI_ISL_73 ···················· ···················· ····················· ···················· ···················· ····················
Y 2540 hCoV-19/Switzerland/VD-ETHZ-320397/2020|EPI_ISL_73 ···················· ···················· ····················· ···················· ···················· ····················
Y 2541 hCoV-19/Switzerland/VD-ETHZ-320399/2020|EPI_ISL_73 ···················· ···················· ····················· ···················· ······K············· ····················
```

| | | | | | | | | |
|---|---|---|---|---|---|---|---|---|
| Y | 2542 | hCoV-19/Switzerland/VD-ETHZ-320400/2020\|EPI_ISL_73 | ···················· | ···················· | ····················· | ···················· | ···················· | ···················· |
| Y | 2543 | hCoV-19/Switzerland/VD-ETHZ-320402/2020\|EPI_ISL_73 | ···················· | ···················· | ····················· | ···················· | ···················· | ···················· |
| Y | 2544 | hCoV-19/Switzerland/VD-ETHZ-320403/2020\|EPI_ISL_73 | ···················· | ···················· | ····················· | ···················· | ···················· | ···················· |
| Y | 2545 | hCoV-19/Switzerland/VD-ETHZ-320404/2020\|EPI_ISL_73 | ···················· | ···················· | ····················· | ···················· | ···················· | ···················· |
| Y | 2547 | hCoV-19/Switzerland/VD-ETHZ-320409/2020\|EPI_ISL_73 | ···················· | ···················· | ····················· | ···················· | ···················· | ···················· |
| Y | 2548 | hCoV-19/Switzerland/VD-ETHZ-320411/2020\|EPI_ISL_73 | ···················· | ···················· | ····················· | ···················· | ···················· | ·············NN····· |
| Y | 2549 | hCoV-19/Switzerland/VD-ETHZ-320413/2020\|EPI_ISL_73 | ···················· | ···················· | ····················· | ···················· | ···················· | ···················· |
| Y | 2550 | hCoV-19/Switzerland/VD-ETHZ-320414/2020\|EPI_ISL_73 | ···················· | ···················· | ····················· | ···················· | ···················· | ···················· |
| Y | 2551 | hCoV-19/Switzerland/VD-ETHZ-320415/2020\|EPI_ISL_73 | ···················· | ···················· | ····················· | ···················· | ···················· | ···················· |
| Y | 2552 | hCoV-19/Switzerland/VD-ETHZ-320416/2020\|EPI_ISL_73 | ···················· | ···················· | ····················· | ···················· | ···················· | ···················· |
| Y | 2553 | hCoV-19/Switzerland/VD-ETHZ-320518/2020\|EPI_ISL_73 | ···················· | ···················· | ····················· | ···················· | ···················· | ···················· |
| Y | 2554 | hCoV-19/Switzerland/VD-ETHZ-320520/2020\|EPI_ISL_73 | ···················· | ···················· | ····················· | ···················· | ···················· | ···················· |
| Y | 2555 | hCoV-19/Switzerland/VD-ETHZ-320521/2020\|EPI_ISL_73 | ···················· | ···················· | ····················· | ···················· | ···················· | ···················· |
| Y | 2556 | hCoV-19/Switzerland/VD-ETHZ-320522/2020\|EPI_ISL_73 | ···················· | ···················· | ····················· | ···················· | ···················· | ···················· |
| Y | 2558 | hCoV-19/Switzerland/VD-ETHZ-320530/2020\|EPI_ISL_73 | ···················· | ···················· | ····················· | ···················· | ···················· | ···················· |
| Y | 2559 | hCoV-19/Switzerland/VD-ETHZ-320531/2020\|EPI_ISL_73 | ···················· | ···················· | ····················· | ···················· | ···················· | ···················· |
| Y | 2560 | hCoV-19/Switzerland/VD-ETHZ-320578/2020\|EPI_ISL_73 | ···················· | ···················· | ····················· | ···················· | ···················· | ···················· |
| Y | 2561 | hCoV-19/England/LOND-1264D59/2020\|EPI_ISL_724563\|2 | ···················· | ···················· | ····················· | ···················· | ·····T·············· | ···················· |
| Y | 2562 | hCoV-19/England/LOND-1264DB3/2020\|EPI_ISL_724567\|2 | ···················· | ·A·················· | ····················· | ···················· | ···················· | ··A················· |
| Y | 2563 | hCoV-19/England/NORW-21F6BF/2020\|EPI_ISL_725040\|20 | ···················· | ···················· | ····················· | ···················· | ···················· | ···················· |
| Y | 2564 | hCoV-19/England/NORW-21F6CE/2020\|EPI_ISL_725041\|20 | ···················· | ·A·················· | ····················· | ···················· | ···················· | ··A················· |
| Y | 2565 | hCoV-19/England/NORW-21F707/2020\|EPI_ISL_725042\|20 | ···················· | ···················· | ····················· | ···················· | ···················· | ···················· |
| Y | 2566 | hCoV-19/England/NORW-21F743/2020\|EPI_ISL_725045\|20 | ···················· | ···················· | ····················· | ···················· | ···················· | ···················· |
| Y | 2567 | hCoV-19/England/NORW-21F752/2020\|EPI_ISL_725046\|20 | ···················· | ···················· | ····················· | ···················· | ···················· | ···················· |
| Y | 2568 | hCoV-19/England/NORW-21F761/2020\|EPI_ISL_725047\|20 | ···················· | ·A·················· | ····················· | ···················· | ···················· | ··A················· |
| Y | 2569 | hCoV-19/England/NORW-21F79E/2020\|EPI_ISL_725049\|20 | ···················· | ···················· | ····················· | ···················· | ···················· | ···················· |
| Y | 2570 | hCoV-19/England/NORW-21F7CB/2020\|EPI_ISL_725050\|20 | ···················· | ···················· | ····················· | ···················· | ···················· | ···················· |
| Y | 2571 | hCoV-19/England/NORW-21F7E9/2020\|EPI_ISL_725051\|20 | ···················· | ···················· | ····················· | ···················· | ···················· | ···················· |
| Y | 2572 | hCoV-19/England/NORW-21F813/2020\|EPI_ISL_725054\|20 | ···················· | ···················· | ····················· | ···················· | ···················· | ···················· |
| Y | 2573 | hCoV-19/England/NORW-21F831/2020\|EPI_ISL_725055\|20 | ···················· | ···················· | ····················· | ···················· | ···················· | ···················· |
| Y | 2574 | hCoV-19/England/NORW-21F840/2020\|EPI_ISL_725056\|20 | ···················· | ···················· | ····················· | ···················· | ···················· | ···················· |
| Y | 2575 | hCoV-19/England/NORW-21F85F/2020\|EPI_ISL_725057\|20 | ···················· | ···················· | ····················· | ···················· | ···················· | ···················· |
| Y | 2576 | hCoV-19/England/NORW-21F87D/2020\|EPI_ISL_725058\|20 | ···················· | ···················· | ····················· | ···················· | ···················· | ···················· |
| Y | 2577 | hCoV-19/England/NORW-21F88C/2020\|EPI_ISL_725059\|20 | ···················· | ···················· | ····················· | ···················· | ···················· | ···················· |
| Y | 2578 | hCoV-19/England/NORW-21F8AA/2020\|EPI_ISL_725060\|20 | ···················· | ···················· | ····················· | ···················· | ···················· | ···················· |
| Y | 2579 | hCoV-19/England/NORW-21F910/2020\|EPI_ISL_725062\|20 | ···················· | ·A·················· | ····················· | ···················· | ···················· | ··A················· |
| Y | 2580 | hCoV-19/England/NORW-21F9C5/2020\|EPI_ISL_725063\|20 | ···················· | ···················· | ····················· | ···················· | ···················· | ···················· |
| Y | 2581 | hCoV-19/England/NORW-21FA1D/2020\|EPI_ISL_725064\|20 | ···················· | ·A·················· | ····················· | ···················· | ···················· | ··A················· |
| Y | 2582 | hCoV-19/England/NORW-21FA68/2020\|EPI_ISL_725066\|20 | ···················· | ···················· | ····················· | ···················· | ···················· | ···················· |
| Y | 2583 | hCoV-19/England/NORW-21FDF6/2020\|EPI_ISL_725068\|20 | ···················· | ···················· | ····················· | ···················· | ···················· | ···················· |
| Y | 2584 | hCoV-19/England/NORW-21FE20/2020\|EPI_ISL_725069\|20 | ···················· | ···················· | ····················· | ···················· | ···················· | ···················· |
| Y | 2585 | hCoV-19/England/NORW-21FE3F/2020\|EPI_ISL_725070\|20 | ···················· | ···················· | ····················· | ···················· | ···················· | ···················· |
| Y | 2586 | hCoV-19/England/NORW-21FE5D/2020\|EPI_ISL_725071\|20 | ···················· | ···················· | ····················· | ···················· | ···················· | ···················· |
| Y | 2587 | hCoV-19/England/NORW-21FE7B/2020\|EPI_ISL_725073\|20 | ···················· | ···················· | ····················· | ···················· | ···················· | ···················· |
| Y | 2588 | hCoV-19/England/NORW-21FE99/2020\|EPI_ISL_725074\|20 | ···················· | ···················· | ····················· | ···················· | ···················· | ···················· |
| Y | 2589 | hCoV-19/England/NORW-21FEC6/2020\|EPI_ISL_725077\|20 | ···················· | ·A·················· | ····················· | ···················· | ···················· | ··A················· |
| Y | 2590 | hCoV-19/England/NORW-21FF1E/2020\|EPI_ISL_725078\|20 | ···················· | ·A·················· | ····················· | ···················· | ···················· | ··A················· |
| Y | 2591 | hCoV-19/Wales/PHWC-49347B/2020\|EPI_ISL_725890\|2020 | ···················· | ···················· | ····················· | ···················· | ···················· | ···················· |
| Y | 2592 | hCoV-19/Wales/PHWC-49348A/2020\|EPI_ISL_725891\|2020 | ···················· | ···················· | ····················· | ···················· | ···················· | ···················· |
| Y | 2594 | hCoV-19/Wales/PHWC-4934D5/2020\|EPI_ISL_725896\|2020 | ···················· | ···················· | ····················· | ···················· | ···················· | ···················· |
| Y | 2595 | hCoV-19/Wales/PHWC-4955EF/2020\|EPI_ISL_726370\|2020 | ···················· | ···················· | ····················· | ···················· | ···················· | ···················· |
| Y | 2598 | hCoV-19/Wales/PHWC-495619/2020\|EPI_ISL_726373\|2020 | ···················· | ···················· | ····················· | ···················· | ···················· | ···················· |
| Y | 2599 | hCoV-19/Wales/PHWC-495637/2020\|EPI_ISL_726374\|2020 | ···················· | ···················· | ····················· | ···················· | ···················· | ···················· |
| Y | 2600 | hCoV-19/Wales/PHWC-495655/2020\|EPI_ISL_726375\|2020 | ···················· | ···················· | ····················· | ···················· | ···················· | ···················· |
| Y | 2601 | hCoV-19/Wales/PHWC-495664/2020\|EPI_ISL_726376\|2020 | ···················· | ···················· | ····················· | ···················· | ···················· | ···················· |
| Y | 2602 | hCoV-19/Wales/PHWC-495682/2020\|EPI_ISL_726378\|2020 | ···················· | ···················· | ····················· | ···················· | ···················· | ···················· |
| Y | 2607 | hCoV-19/Wales/PHWC-495734/2020\|EPI_ISL_726389\|2020 | ···················· | ···················· | ····················· | ···················· | ···················· | ···················· |
| Y | 2608 | hCoV-19/Wales/PHWC-495743/2020\|EPI_ISL_726390\|2020 | ···················· | ···················· | ····················· | ···················· | ···················· | ···················· |
| Y | 2609 | hCoV-19/Wales/PHWC-495752/2020\|EPI_ISL_726391\|2020 | ···················· | ···················· | ····················· | ···················· | ···················· | ···················· |
| Y | 2610 | hCoV-19/Wales/PHWC-495761/2020\|EPI_ISL_726392\|2020 | ···················· | ···················· | ····················· | ···················· | ···················· | ···················· |
| Y | 2611 | hCoV-19/Wales/PHWC-495770/2020\|EPI_ISL_726393\|2020 | ···················· | ···················· | ····················· | ···················· | ···················· | ···················· |
| Y | 2613 | hCoV-19/Wales/PHWC-4957AD/2020\|EPI_ISL_726395\|2020 | ···················· | ···················· | ····················· | ···················· | ···················· | ···················· |
| Y | 2614 | hCoV-19/Wales/PHWC-4957BC/2020\|EPI_ISL_726396\|2020 | ···················· | ···················· | ····················· | ···················· | ···················· | ···················· |
| Y | 2616 | hCoV-19/Wales/PHWC-4957DA/2020\|EPI_ISL_726398\|2020 | ···················· | ···················· | ····················· | ···················· | ···················· | ···················· |
| Y | 2617 | hCoV-19/England/ALDP-C3BC39/2020\|EPI_ISL_731700\|20 | ···················· | ···················· | ····················· | ···················· | ···················· | ···················· |
| Y | 2618 | hCoV-19/England/ALDP-C3B088/2020\|EPI_ISL_731701\|20 | ···················· | ···················· | ····················· | ···················· | ···················· | ···················· |
| Y | 2619 | hCoV-19/England/ALDP-C3CB0E/2020\|EPI_ISL_731702\|20 | ···················· | ···················· | ····················· | ···················· | ···················· | ···················· |
| Y | 2620 | hCoV-19/England/ALDP-C3BC66/2020\|EPI_ISL_731704\|20 | ···················· | ·A·················· | ····················· | ···················· | ···················· | ···················· |
| Y | 2621 | hCoV-19/England/ALDP-C3CA01/2020\|EPI_ISL_731705\|20 | ···················· | ···················· | ····················· | ···················· | ···················· | ···················· |
| Y | 2622 | hCoV-19/England/ALDP-C3D2DB/2020\|EPI_ISL_731706\|20 | ···················· | ···················· | ····················· | ···················· | ···················· | ···················· |
| Y | 2623 | hCoV-19/England/ALDP-C3B01F/2020\|EPI_ISL_731707\|20 | ···················· | ···················· | ····················· | ···················· | ···················· | ···················· |
| Y | 2624 | hCoV-19/England/ALDP-C3C737/2020\|EPI_ISL_731708\|20 | ···················· | ···················· | ····················· | ···················· | ···················· | ···················· |
| Y | 2625 | hCoV-19/England/ALDP-C3B307/2020\|EPI_ISL_731709\|20 | ···················· | ···················· | ····················· | ···················· | ···················· | ···················· |
| Y | 2626 | hCoV-19/England/ALDP-C3D3D8/2020\|EPI_ISL_731710\|20 | ···················· | ···················· | ····················· | ···················· | ···················· | ···················· |
| Y | 2627 | hCoV-19/Belgium/rega-0720809/2020\|EPI_ISL_734802\|2 | ···················· | ···················· | ····················· | ···················· | ···················· | ···················· |
| Y | 2628 | hCoV-19/Belgium/rega-0720810/2020\|EPI_ISL_734803\|2 | ···················· | ···················· | ····················· | ···················· | ···················· | ···················· |
| Y | 2629 | hCoV-19/Belgium/rega-0721813/2020\|EPI_ISL_734806\|2 | ···················· | ···················· | ····················· | ···················· | ···················· | ···················· |
| Y | 2630 | hCoV-19/Belgium/rega-0721837/2020\|EPI_ISL_734807\|2 | ···················· | ···················· | ····················· | ···················· | ···················· | ···················· |
| Y | 2631 | hCoV-19/Belgium/rega-0721838/2020\|EPI_ISL_734808\|2 | ···················· | ···················· | ····················· | ···················· | ···················· | ···················· |
| Y | 2632 | hCoV-19/Belgium/rega-0721839/2020\|EPI_ISL_734809\|2 | ···················· | ···················· | ····················· | ···················· | ···················· | ···················· |

| | | | | | | | |
|---|---|---|---|---|---|---|---|
| Y | 2633 hCoV-19/Belgium/rega-0721843/2020\|EPI_ISL_734813\|2 | .................... | .................... | ..................... | .................... | .................... | .................... |
| Y | 2634 hCoV-19/Belgium/rega-0721844/2020\|EPI_ISL_734814\|2 | .................... | .................... | ..................... | .................... | .................... | .................... |
| Y | 2635 hCoV-19/Belgium/rega-0722802/2020\|EPI_ISL_734815\|2 | .................... | .................... | ..................... | .................... | .................... | .................... |
| Y | 2636 hCoV-19/Belgium/rega-0722803/2020\|EPI_ISL_734816\|2 | .................... | .................... | ..................... | .................... | .................... | .................... |
| Y | 2637 hCoV-19/Belgium/rega-0722804/2020\|EPI_ISL_734817\|2 | .................... | .................... | ..................... | .................... | .................... | .................... |
| Y | 2638 hCoV-19/Belgium/rega-0722814/2020\|EPI_ISL_734818\|2 | .................... | .................... | ..................... | .................... | .................... | .................... |
| Y | 2639 hCoV-19/Belgium/rega-0722815/2020\|EPI_ISL_734819\|2 | .................... | .................... | ..................... | .................... | .................... | .................... |
| Y | 2640 hCoV-19/Belgium/rega-0722816/2020\|EPI_ISL_734820\|2 | .................... | .................... | ..................... | .................... | .................... | .................... |
| Y | 2641 hCoV-19/Belgium/rega-0722817/2020\|EPI_ISL_734821\|2 | .................... | .................... | ..................... | .................... | .................... | .................... |
| Y | 2642 hCoV-19/Belgium/rega-0722818/2020\|EPI_ISL_734822\|2 | .................... | .................... | ..................... | .................... | .................... | .................... |
| Y | 2643 hCoV-19/Belgium/rega-0722819/2020\|EPI_ISL_734823\|2 | .................... | .................... | ..................... | .................... | .................... | .................... |
| Y | 2644 hCoV-19/Belgium/rega-10191296/2020\|EPI_ISL_735200\| | .................... | .................... | ..................... | .................... | .................... | .................... |
| Y | 2645 hCoV-19/Belgium/rega-10191297/2020\|EPI_ISL_735201\| | .................... | .................... | ..................... | .................... | .................... | .................... |
| Y | 2646 hCoV-19/Belgium/rega-10191298/2020\|EPI_ISL_735202\| | .................... | .................... | ..................... | .................... | .................... | .................... |
| Y | 2647 hCoV-19/Belgium/rega-10191299/2020\|EPI_ISL_735203\| | .................... | .................... | ..................... | .................... | .................... | .................... |
| Y | 2648 hCoV-19/Belgium/rega-10191301/2020\|EPI_ISL_735204\| | .................... | .................... | ..................... | .................... | .................... | .................... |
| Y | 2649 hCoV-19/Belgium/rega-10191302/2020\|EPI_ISL_735205\| | .................... | .................... | ..................... | .................... | .................... | .................... |
| Y | 2650 hCoV-19/Belgium/rega-10191303/2020\|EPI_ISL_735206\| | .................... | .................... | ..................... | .................... | .................... | .................... |
| Y | 2651 hCoV-19/Belgium/rega-10191304/2020\|EPI_ISL_735207\| | .................... | .................... | ..................... | .................... | .................... | .................... |
| Y | 2653 hCoV-19/Belgium/rega-12011202/2020\|EPI_ISL_735240\| | .................... | .................... | ..................... | .................... | .................... | .................... |
| Y | 2654 hCoV-19/France/HDF-rega-0413575/2020\|EPI_ISL_73524 | .................... | .................... | ..................... | .................... | .................... | .................... |
| Y | 2655 hCoV-19/France/HDF-rega-0414581/2020\|EPI_ISL_73524 | .................... | .................... | ..................... | .................... | .................... | .................... |
| Y | 2656 hCoV-19/France/HDF-rega-0417506/2020\|EPI_ISL_73524 | .................... | .................... | ..................... | .................... | .................... | .................... |
| Y | 2657 hCoV-19/France/NOR-rega-0423601/2020\|EPI_ISL_73524 | .................... | .................... | ..................... | .................... | .................... | .................... |
| Y | 2658 hCoV-19/France/IDF-rega-0508543/2020\|EPI_ISL_73524 | .................... | .................... | ..................... | .................... | .................... | .................... |
| Y | 2659 hCoV-19/France/HDF-rega-0510548/2020\|EPI_ISL_73524 | .................... | .................... | ..................... | .................... | .................... | .................... |
| Y | 2660 hCoV-19/France/HDF-rega-0703808/2020\|EPI_ISL_73524 | .................... | .................... | ..................... | .................... | .................... | .................... |
| Y | 2661 hCoV-19/Netherlands/rega-0426602/2020\|EPI_ISL_7352 | .................... | .................... | ..................... | .................... | .................... | .................... |
| Y | 2662 hCoV-19/Belgium/rega-0429664/2020\|EPI_ISL_735249\|2 | .................... | .................... | ..................... | .................... | .................... | .................... |
| Y | 2663 hCoV-19/Turkey/IST-GLAB-010/2020\|EPI_ISL_735260\|20 | .................... | .................... | ..................... | .................... | .................... | .................... |
| Y | 2664 hCoV-19/Turkey/IST-GLAB-026/2020\|EPI_ISL_735261\|20 | .................... | .................... | ..................... | .................... | .................... | .................... |
| Y | 2665 hCoV-19/Turkey/IST-GLAB-028/2020\|EPI_ISL_735262\|20 | .................... | .................... | ..................... | .................... | .................... | .................... |
| Y | 2666 hCoV-19/Turkey/IST-GLAB-056/2020\|EPI_ISL_735263\|20 | .................... | .................... | ..................... | .................... | .................... | .................... |
| Y | 2667 hCoV-19/Turkey/IST-GLAB-060/2020\|EPI_ISL_735264\|20 | .................... | .................... | ..................... | .................... | .................... | .................... |
| Y | 2668 hCoV-19/Turkey/IST-GLAB-061/2020\|EPI_ISL_735265\|20 | .................... | .................... | ..................... | .................... | .................... | .................... |
| Y | 2669 hCoV-19/Turkey/IST-GLAB-065/2020\|EPI_ISL_735266\|20 | .................... | .................... | ..................... | .................... | .................... | .................... |
| Y | 2670 hCoV-19/Turkey/IST-GLAB-119/2020\|EPI_ISL_735267\|20 | .................... | .................... | ..................... | .................... | .................... | .................... |
| Y | 2671 hCoV-19/Turkey/IST-GLAB-120/2020\|EPI_ISL_735268\|20 | .................... | .................... | ..................... | .................... | .................... | .................... |
| Y | 2672 hCoV-19/Turkey/IST-GLAB-121/2020\|EPI_ISL_735269\|20 | .................... | .................... | ..................... | .................... | .................... | .................... |
| Y | 2673 hCoV-19/Turkey/IST-GLAB-122/2020\|EPI_ISL_735270\|20 | .................... | .................... | ..................... | .................... | .................... | .................... |
| Y | 2674 hCoV-19/Turkey/IST-GLAB-123/2020\|EPI_ISL_735271\|20 | .................... | .................... | ..................... | .................... | .................... | .................... |
| Y | 2675 hCoV-19/Turkey/IST-GLAB-124/2020\|EPI_ISL_735272\|20 | .................... | .................... | ..................... | .................... | .................... | .................... |
| Y | 2676 hCoV-19/Turkey/IST-GLAB-127/2020\|EPI_ISL_735273\|20 | .................... | .................... | ..................... | .................... | .................... | .................... |
| Y | 2677 hCoV-19/Turkey/IST-GLAB-128/2020\|EPI_ISL_735274\|20 | .................... | .................... | ..................... | .................... | .................... | .................... |
| Y | 2678 hCoV-19/Turkey/IST-GLAB-129/2020\|EPI_ISL_735275\|20 | .................... | .................... | ..................... | .................... | .................... | .................... |
| Y | 2679 hCoV-19/Turkey/IST-GLAB-131/2020\|EPI_ISL_735276\|20 | .................... | .................... | ..................... | .................... | .................... | .................... |
| Y | 2680 hCoV-19/Turkey/IST-GLAB-140/2020\|EPI_ISL_735277\|20 | .................... | .................... | ..................... | .................... | .................... | .................... |
| Y | 2681 hCoV-19/Turkey/IST-GLAB-141/2020\|EPI_ISL_735278\|20 | .................... | .................... | ..................... | .................... | .................... | .................... |
| Y | 2682 hCoV-19/Turkey/IST-GLAB-142/2020\|EPI_ISL_735279\|20 | .................... | .................... | ..................... | .................... | .................... | .................... |
| Y | 2683 hCoV-19/Turkey/IST-GLAB-143/2020\|EPI_ISL_735280\|20 | .................... | .................... | ..................... | .................... | .................... | .................... |
| Y | 2684 hCoV-19/Turkey/IST-GLAB-145/2020\|EPI_ISL_735281\|20 | .................... | .................... | ..................... | .................... | .................... | .................... |
| Y | 2685 hCoV-19/Turkey/IST-GLAB-146/2020\|EPI_ISL_735282\|20 | .................... | .................... | ..................... | .................... | .................... | .................... |
| Y | 2686 hCoV-19/Turkey/IST-GLAB-148/2020\|EPI_ISL_735283\|20 | .................... | .................... | ..................... | .................... | .................... | .................... |
| Y | 2687 hCoV-19/Turkey/IST-GLAB-151/2020\|EPI_ISL_735284\|20 | .................... | .................... | ..................... | .................... | .................... | .................... |
| Y | 2688 hCoV-19/Turkey/IST-GLAB-160/2020\|EPI_ISL_735285\|20 | .................... | .................... | ..................... | .................... | .................... | .................... |
| Y | 2689 hCoV-19/Turkey/IST-GLAB-172/2020\|EPI_ISL_735286\|20 | .................... | .................... | ..................... | .................... | .................... | .................... |
| Y | 2690 hCoV-19/Turkey/IST-GLAB-218/2020\|EPI_ISL_735288\|20 | .................... | .A.................. | ..................... | .................... | .................... | ...A................ |
| Y | 2691 hCoV-19/Turkey/IST-GLAB-219/2020\|EPI_ISL_735289\|20 | .................... | .................... | ..................... | .................... | .................... | .................... |
| Y | 2692 hCoV-19/Turkey/IST-GLAB-222/2020\|EPI_ISL_735290\|20 | .................... | .................... | ..................... | .................... | .................... | .................... |
| Y | 2693 hCoV-19/Turkey/IST-GLAB-224/2020\|EPI_ISL_735292\|20 | .................... | ..........A......... | ..................... | .................... | .................... | .................... |
| Y | 2694 hCoV-19/Turkey/IST-GLAB-225/2020\|EPI_ISL_735293\|20 | .................... | .................... | ..................... | .................... | .................... | .................... |
| Y | 2695 hCoV-19/Turkey/IST-GLAB-227/2020\|EPI_ISL_735294\|20 | .................... | ....G............... | ..................... | .................... | .................... | .................... |
| Y | 2696 hCoV-19/Turkey/IST-GLAB-228/2020\|EPI_ISL_735295\|20 | .................... | .................... | ..................... | .................... | .................... | .................... |
| Y | 2697 hCoV-19/Turkey/IST-GLAB-229/2020\|EPI_ISL_735296\|20 | .................... | .................... | ..................... | .................... | .................... | .................... |
| Y | 2698 hCoV-19/Turkey/IST-GLAB-230/2020\|EPI_ISL_735297\|20 | .................... | .................... | ..................... | .................... | .................... | .................... |
| Y | 2699 hCoV-19/Turkey/IST-GLAB-231/2020\|EPI_ISL_735298\|20 | .................... | .................... | ..................... | .................... | .................... | .................... |
| Y | 2700 hCoV-19/Turkey/IST-GLAB-232/2020\|EPI_ISL_735299\|20 | .................... | .................... | ..................... | .................... | .................... | .................... |
| Y | 2701 hCoV-19/England/QEUH-C4BB84/2020\|EPI_ISL_736532\|20 | .................... | .................... | ..................... | .................... | .................... | .................... |
| Y | 2702 hCoV-19/England/QEUH-C4BD51/2020\|EPI_ISL_736533\|20 | .................... | .A.................. | ..................... | .................... | .................... | ...A................ |
| Y | 2703 hCoV-19/England/ALDP-C48E06/2020\|EPI_ISL_736537\|20 | .................... | .................... | ..................... | .................... | .................... | .................... |
| Y | 2704 hCoV-19/England/ALDP-C486A4/2020\|EPI_ISL_736539\|20 | .................... | .................... | ..................... | .................... | .................... | .................... |
| Y | 2705 hCoV-19/England/ALDP-C4863B/2020\|EPI_ISL_736570\|20 | .................... | .................... | ..................... | .................... | .................... | .................... |
| Y | 2706 hCoV-19/England/ALDP-C48A3F/2020\|EPI_ISL_736574\|20 | .................... | .................... | ..................... | .................... | .................... | .................... |
| Y | 2707 hCoV-19/England/ALDP-C489AB/2020\|EPI_ISL_736575\|20 | .................... | .................... | ..................... | .................... | .................... | .................... |
| Y | 2708 hCoV-19/England/ALDP-C48AD5/2020\|EPI_ISL_736577\|20 | .................... | .A.................. | ..................... | .................... | .................... | ...A................ |
| Y | 2709 hCoV-19/England/ALDP-C48D18/2020\|EPI_ISL_736579\|20 | .................... | .................... | ..................... | .................... | .................... | .................... |
| Y | 2710 hCoV-19/England/QEUH-C4BDAC/2020\|EPI_ISL_736580\|20 | .................... | .................... | ..................... | .................... | .................... | .................... |
| Y | 2711 hCoV-19/England/QEUH-C4BDE8/2020\|EPI_ISL_736582\|20 | .................... | .................... | ..................... | .................... | .................... | .................... |
| Y | 2712 hCoV-19/England/ALDP-C4857A/2020\|EPI_ISL_736583\|20 | .................... | .................... | ..................... | .................... | ......T............. | .................... |
| Y | 2713 hCoV-19/England/ALDP-C4899C/2020\|EPI_ISL_736585\|20 | .................... | .................... | ..................... | .................... | .................... | .................... |

| | | | | | | | | |
|---|---|---|---|---|---|---|---|---|
| Y | 2714 hCoV-19/England/ALDP-C489E7/2020\|EPI_ISL_736587\|20 | ···················· | ···················· | ···················· | ···················· | ···················· | ···················· | |
| Y | 2715 hCoV-19/England/MILK-C508A3/2020\|EPI_ISL_736590\|20 | ···················· | ···················· | ···················· | ···················· | ···················· | ···················· | |
| Y | 2716 hCoV-19/England/MILK-C505CA/2020\|EPI_ISL_736597\|20 | ···················· | ···················· | ···················· | ···················· | ···················· | ···················· | |
| Y | 2717 hCoV-19/England/ALDP-C48B1E/2020\|EPI_ISL_736599\|20 | ···················· | ···················· | ···················· | ···················· | ···················· | ···················· | |
| Y | 2718 hCoV-19/Czech Republic/NRL_10961/2020\|EPI_ISL_7370 | ···················· | ···················· | ···················· | ···················· | ···················· | ···················· | |
| Y | 2719 hCoV-19/Czech Republic/NRL_10980/2020\|EPI_ISL_7370 | ···················· | ···················· | ···················· | ···················· | ···················· | ···················· | |
| Y | 2720 hCoV-19/Czech Republic/NRL_10885/2020\|EPI_ISL_7370 | ···················· | ···················· | ···················· | ···················· | ···················· | ···················· | |
| Y | 2721 hCoV-19/Czech Republic/NRL_10888/2020\|EPI_ISL_7370 | ···················· | ···················· | ···················· | ···················· | ···················· | ···················· | |
| Y | 2722 hCoV-19/Czech Republic/NRL_10981/2020\|EPI_ISL_7370 | ···················· | ···················· | ···················· | ···················· | ···················· | ···················· | |
| Y | 2723 hCoV-19/Czech Republic/NRL_10985/2020\|EPI_ISL_7370 | ···················· | ···················· | ···················· | ···················· | ···················· | ···················· | |
| Y | 2724 hCoV-19/Czech Republic/NRL_11016/2020\|EPI_ISL_7370 | ···················· | ···················· | ···················· | ···················· | ···················· | ···················· | |
| Y | 2725 hCoV-19/Czech Republic/NRL_11017/2020\|EPI_ISL_7370 | ···················· | ···················· | ···················· | ···················· | ···················· | ···················· | |
| Y | 2726 hCoV-19/Czech Republic/NRL_10571/2020\|EPI_ISL_7370 | ···················· | ···················· | ···················· | ···················· | ···················· | ···················· | |
| Y | 2727 hCoV-19/Czech Republic/NRL_10533/2020\|EPI_ISL_7370 | ···················· | ···················· | ···················· | ···················· | ···················· | ···················· | |
| Y | 2728 hCoV-19/Czech Republic/NRL_10569/2020\|EPI_ISL_7370 | ···················· | ···················· | ···················· | ···················· | ···················· | ···················· | |
| Y | 2729 hCoV-19/Czech Republic/NRL_10593/2020\|EPI_ISL_7370 | ···················· | ···················· | ···················· | ···················· | ···················· | ···················· | |
| Y | 2730 hCoV-19/Czech Republic/NRL_10594/2020\|EPI_ISL_7370 | ···················· | ···················· | ···················· | ···················· | ···················· | ···················· | |
| Y | 2731 hCoV-19/Finland/S10E4/2020\|EPI_ISL_737034\|2020-08- | ···················· | ·A·················· | ···················· | ···················· | ···················· | ··A················· | |
| Y | 2732 hCoV-19/Finland/S1B4/2020\|EPI_ISL_737035\|2020-08-1 | ···················· | ···················· | ···················· | ···················· | ···················· | ···················· | |
| Y | 2733 hCoV-19/Finland/S24C2/2020\|EPI_ISL_737036\|2020-08- | ···················· | ···················· | ···················· | ···················· | ···················· | ···················· | |
| Y | 2734 hCoV-19/Finland/S10AE4/2020\|EPI_ISL_737037\|2020-10 | ···················· | ·A·················· | ···················· | ···················· | ···················· | ··A················· | |
| Y | 2735 hCoV-19/Finland/S10AG10/2020\|EPI_ISL_737038\|2020-1 | ···················· | ···················· | ···················· | ···················· | ···················· | ···················· | |
| Y | 2736 hCoV-19/Finland/S10AH10/2020\|EPI_ISL_737039\|2020-1 | ···················· | ·A·················· | ···················· | ···················· | ···················· | ··A················· | |
| Y | 2737 hCoV-19/Finland/S10AH9/2020\|EPI_ISL_737040\|2020-10 | ···················· | ···················· | ···················· | ···················· | ···················· | ···················· | |
| Y | 2738 hCoV-19/Finland/S12AH3/2020\|EPI_ISL_737041\|2020-10 | ···················· | ·A·················· | ···················· | ···················· | ···················· | ··A················· | |
| Y | 2739 hCoV-19/Finland/S13AD3/2020\|EPI_ISL_737042\|2020-10 | ···················· | ···················· | ···················· | ···················· | ···················· | ···················· | |
| Y | 2740 hCoV-19/Finland/S13AD9/2020\|EPI_ISL_737043\|2020-10 | ···················· | ···················· | ···················· | ···················· | ···················· | ···················· | |
| Y | 2741 hCoV-19/Finland/S2BA11/2020\|EPI_ISL_737044\|2020-10 | ···················· | ···················· | ···················· | ···················· | ···················· | ···················· | |
| Y | 2742 hCoV-19/Finland/S4AA5/2020\|EPI_ISL_737045\|2020-10- | ···················· | ···················· | ···················· | ···················· | ···················· | ···················· | |
| Y | 2743 hCoV-19/Finland/S4AB3/2020\|EPI_ISL_737046\|2020-10- | ···················· | ···················· | ···················· | ···················· | ···················· | ···················· | |
| Y | 2744 hCoV-19/Finland/S4AD5/2020\|EPI_ISL_737047\|2020-10- | ···················· | ···················· | ···················· | ···················· | ···················· | ···················· | |
| Y | 2745 hCoV-19/Finland/S4BD9/2020\|EPI_ISL_737048\|2020-10- | ···················· | ·A·················· | ···················· | ···················· | ···················· | ··A················· | |
| Y | 2746 hCoV-19/Finland/S4BG9/2020\|EPI_ISL_737049\|2020-10- | ···················· | ·A·················· | ···················· | ···················· | ···················· | ··A················· | |
| Y | 2747 hCoV-19/Finland/S6AA10/2020\|EPI_ISL_737050\|2020-10 | ···················· | ·A·················· | ···················· | ···················· | ···················· | ··A················· | |
| Y | 2748 hCoV-19/Finland/S6AB2/2020\|EPI_ISL_737051\|2020-10- | ···················· | ···················· | ···················· | ···················· | ···················· | ···················· | |
| Y | 2749 hCoV-19/Finland/S6AC11/2020\|EPI_ISL_737052\|2020-10 | ···················· | ···················· | ···················· | ···················· | ···················· | ···················· | |
| Y | 2750 hCoV-19/Finland/S6AH10/2020\|EPI_ISL_737053\|2020-10 | ···················· | ···················· | ···················· | ···················· | ···················· | ···················· | |
| Y | 2751 hCoV-19/Finland/S7AA11/2020\|EPI_ISL_737054\|2020-10 | ···················· | ···················· | ···················· | ···················· | ···················· | ···················· | |
| Y | 2752 hCoV-19/Finland/S8AA11/2020\|EPI_ISL_737055\|2020-10 | ···················· | ·A·················· | ···················· | ···················· | ···················· | ··A················· | |
| Y | 2753 hCoV-19/Finland/S8AC11/2020\|EPI_ISL_737056\|2020-10 | ···················· | ·A·················· | ···················· | ···················· | ···················· | ··A················· | |
| Y | 2754 hCoV-19/Finland/S8AC7/2020\|EPI_ISL_737057\|2020-10- | ···················· | ·A·················· | ···················· | ···················· | ···················· | ··A················· | |
| Y | 2755 hCoV-19/Finland/S8AE4/2020\|EPI_ISL_737058\|2020-10- | ···················· | ·A·················· | ···················· | ···················· | ···················· | ··A················· | |
| Y | 2756 hCoV-19/Finland/S8AG2/2020\|EPI_ISL_737059\|2020-10- | ···················· | ···················· | ···················· | ···················· | ···················· | ···················· | |
| Y | 2757 hCoV-19/Finland/S9A4/2020\|EPI_ISL_737080\|2020-10-1 | ···················· | ·A·················· | ···················· | ···················· | ···················· | ··A················· | |
| Y | 2758 hCoV-19/USA/WA-UW-32725/2020\|EPI_ISL_737081\|2020-1 | ···················· | ·A·················· | ···················· | ···················· | ···················· | ···················· | |
| Y | 2759 hCoV-19/USA/WA-UW-32735/2020\|EPI_ISL_737083\|2020-1 | ···················· | ·A·················· | ···················· | ···················· | ···················· | ···················· | |
| Y | 2760 hCoV-19/USA/WA-UW-32746/2020\|EPI_ISL_737084\|2020-1 | ···················· | ···················· | ···················· | ···················· | ···················· | ···················· | |
| Y | 2761 hCoV-19/USA/WA-UW-32820/2020\|EPI_ISL_737090\|2020-1 | ···················· | ···················· | ···················· | ···················· | ···················· | ···················· | |
| Y | 2762 hCoV-19/USA/WA-UW-32824/2020\|EPI_ISL_737091\|2020-1 | ···················· | ···················· | ···················· | ···················· | ···················· | ··DD·T·············· | 1 |
| Y | 2763 hCoV-19/USA/WA-UW-32830/2020\|EPI_ISL_737092\|2020-1 | ···················· | ·A·················· | ···················· | ···················· | ···················· | ···················· | |
| Y | 2764 hCoV-19/USA/WA-UW-32886/2020\|EPI_ISL_737094\|2020-1 | ···················· | ···················· | ···················· | ···················· | ···················· | ···················· | |
| Y | 2765 hCoV-19/USA/WA-UW-32893/2020\|EPI_ISL_737095\|2020-1 | ···················· | ·A·················· | ···················· | ···················· | ···················· | ···················· | |
| Y | 2766 hCoV-19/USA/WA-UW-32904/2020\|EPI_ISL_737096\|2020-1 | ···················· | ·A·················· | ···················· | ···················· | ···················· | ···················· | |
| Y | 2767 hCoV-19/USA/WA-UW-32910/2020\|EPI_ISL_737097\|2020-1 | ···················· | ·A·················· | ···················· | ···················· | ···················· | ···················· | |
| Y | 2768 hCoV-19/USA/WA-UW-32915/2020\|EPI_ISL_737098\|2020-1 | ···················· | ·A·················· | ···················· | ···················· | ···················· | ···················· | |
| Y | 2769 hCoV-19/Switzerland/VS-ETHZ-410638/2020\|EPI_ISL_73 | ···················· | ···················· | ···················· | ···················· | ···················· | ···················· | |
| Y | 2770 hCoV-19/Switzerland/VS-ETHZ-410653/2020\|EPI_ISL_73 | ···················· | ···················· | ···················· | ···················· | ···················· | ···················· | |
| Y | 2771 hCoV-19/Switzerland/VS-ETHZ-410679/2020\|EPI_ISL_73 | ···················· | ···················· | ···················· | ···················· | ···················· | ···················· | |
| Y | 2772 hCoV-19/Switzerland/VS-ETHZ-410723/2020\|EPI_ISL_73 | ···················· | ···················· | ···················· | ···················· | ···················· | ···················· | |
| Y | 2773 hCoV-19/Switzerland/VS-ETHZ-410736/2020\|EPI_ISL_73 | ···················· | ···················· | ···················· | ···················· | ···················· | ···················· | |
| Y | 2774 hCoV-19/Switzerland/VS-ETHZ-410737/2020\|EPI_ISL_73 | ···················· | ···················· | ···················· | ···················· | ···················· | ···················· | |
| Y | 2775 hCoV-19/Switzerland/VS-ETHZ-410769/2020\|EPI_ISL_73 | ···················· | ···················· | ···················· | ···················· | ···················· | ···················· | |
| Y | 2776 hCoV-19/Switzerland/VS-ETHZ-410808/2020\|EPI_ISL_73 | ···················· | ···················· | ···················· | ···················· | ···················· | ···················· | |
| Y | 2777 hCoV-19/Switzerland/VS-ETHZ-410824/2020\|EPI_ISL_73 | ···················· | ···················· | ···················· | ···················· | ···················· | ···················· | |
| Y | 2778 hCoV-19/Switzerland/VS-ETHZ-410870/2020\|EPI_ISL_73 | ···················· | ···················· | ···················· | ···················· | ···················· | ···················· | |
| Y | 2779 hCoV-19/USA/UT-UPHL-2012522014/2020\|EPI_ISL_738352 | ···················· | ···················· | ···················· | ···················· | ···················· | ···················· | |
| Y | 2780 hCoV-19/USA/UT-UPHL-2012039803/2020\|EPI_ISL_738353 | ···················· | ···················· | ···················· | ···················· | ···················· | ···················· | |
| Y | 2781 hCoV-19/USA/UT-UPHL-2012983373/2020\|EPI_ISL_738354 | ···················· | ···················· | ···················· | ···················· | ···················· | ···················· | |
| Y | 2782 hCoV-19/USA/UT-UPHL-2012566145/2020\|EPI_ISL_738355 | ···················· | ···················· | ···················· | ···················· | ···················· | ···················· | |
| Y | 2783 hCoV-19/USA/UT-UPHL-2012045106/2020\|EPI_ISL_738356 | ···················· | ···················· | ···················· | ···················· | ···················· | ···················· | |
| Y | 2784 hCoV-19/USA/UT-UPHL-2012558654/2020\|EPI_ISL_738357 | ···················· | ···················· | ···················· | ···················· | ···················· | ···················· | |
| Y | 2785 hCoV-19/USA/UT-UPHL-2012218768/2020\|EPI_ISL_738358 | ···················· | ···················· | ···················· | ···················· | ···················· | ···················· | |
| Y | 2786 hCoV-19/USA/UT-UPHL-2012875547/2020\|EPI_ISL_738359 | ···················· | ···················· | ···················· | ···················· | ···················· | ···················· | |
| Y | 2787 hCoV-19/Belgium/rega-10141378/2020\|EPI_ISL_738360\| | ···················· | ···················· | ···················· | ···················· | ···················· | ···················· | |
| Y | 2788 hCoV-19/Belgium/rega-12091307/2020\|EPI_ISL_738361\| | ···················· | ···················· | ···················· | ···················· | ···················· | ···················· | |
| Y | 2789 hCoV-19/Belgium/rega-12111308/2020\|EPI_ISL_738362\| | ···················· | ···················· | ···················· | ···················· | ···················· | ···················· | |
| Y | 2790 hCoV-19/Belgium/rega-12141309/2020\|EPI_ISL_738363\| | ···················· | ···················· | ···················· | ···················· | ···················· | ···················· | |
| Y | 2791 hCoV-19/Belgium/rega-12111310/2020\|EPI_ISL_738364\| | ···················· | ···················· | ···················· | ···················· | ···················· | ···················· | |
| Y | 2792 hCoV-19/Belgium/rega-12141311/2020\|EPI_ISL_738365\| | ···················· | ···················· | ···················· | ···················· | ···················· | ···················· | |
| Y | 2793 hCoV-19/Belgium/rega-12111312/2020\|EPI_ISL_738366\| | ···················· | ···················· | ···················· | ···················· | ···················· | ···················· | |

| | | | | | | | |
|---|---|---|---|---|---|---|---|
| Y | 2794 hCoV-19/Belgium/rega-12111313/2020\|EPI_ISL_738367\| | ···················· | ···················· | ····················· | ···················· | ···················· | ···················· |
| Y | 2795 hCoV-19/Belgium/rega-12111314/2020\|EPI_ISL_738368\| | ···················· | ···················· | ····················· | ···················· | ···················· | ···················· |
| Y | 2796 hCoV-19/Belgium/rega-12141315/2020\|EPI_ISL_738369\| | ···················· | ···················· | ····················· | ···················· | ···················· | ···················· |
| Y | 2797 hCoV-19/Belgium/rega-12111316/2020\|EPI_ISL_738370\| | ···················· | ···················· | ····················· | ···················· | ···················· | ···················· |
| Y | 2798 hCoV-19/Belgium/rega-12081318/2020\|EPI_ISL_738371\| | ···················· | ···················· | ····················· | ···················· | ···················· | ···················· |
| Y | 2799 hCoV-19/Belgium/rega-12111319/2020\|EPI_ISL_738372\| | ···················· | ···················· | ····················· | ···················· | ···················· | ···················· |
| Y | 2800 hCoV-19/Belgium/rega-12111320/2020\|EPI_ISL_738373\| | ···················· | ···················· | ····················· | ···················· | ···················· | ···················· |
| Y | 2801 hCoV-19/Belgium/rega-12141321/2020\|EPI_ISL_738374\| | ···················· | ···················· | ····················· | ···················· | ···················· | ···················· |
| Y | 2802 hCoV-19/Belgium/rega-12111322/2020\|EPI_ISL_738375\| | ···················· | ···················· | ····················· | ···················· | ···················· | ···················· |
| Y | 2803 hCoV-19/Belgium/rega-12141323/2020\|EPI_ISL_738376\| | ···················· | ···················· | ····················· | ···················· | ···················· | ···················· |
| Y | 2804 hCoV-19/Belgium/rega-12111324/2020\|EPI_ISL_738377\| | ···················· | ···················· | ····················· | ···················· | ···················· | ···················· |
| Y | 2805 hCoV-19/Belgium/rega-12111325/2020\|EPI_ISL_738378\| | ···················· | ···················· | ····················· | ···················· | ···················· | ···················· |
| Y | 2806 hCoV-19/Belgium/rega-12111326/2020\|EPI_ISL_738379\| | ···················· | ···················· | ····················· | ···················· | ···················· | ···················· |
| Y | 2807 hCoV-19/Belgium/rega-12141327/2020\|EPI_ISL_738380\| | ···················· | ···················· | ····················· | ···················· | ···················· | ···················· |
| Y | 2808 hCoV-19/Belgium/rega-12141328/2020\|EPI_ISL_738381\| | ···················· | ···················· | ····················· | ···················· | ···················· | ···················· |
| Y | 2809 hCoV-19/Belgium/rega-12141329/2020\|EPI_ISL_738382\| | ···················· | ···················· | ····················· | ···················· | ···················· | ···················· |
| Y | 2810 hCoV-19/Belgium/rega-12111330/2020\|EPI_ISL_738383\| | ···················· | ···················· | ····················· | ···················· | ···················· | ···················· |
| Y | 2811 hCoV-19/Belgium/rega-12111331/2020\|EPI_ISL_738384\| | ···················· | ···················· | ····················· | ···················· | ···················· | ···················· |
| Y | 2812 hCoV-19/Belgium/rega-12111332/2020\|EPI_ISL_738385\| | ···················· | ···················· | ····················· | ···················· | ···················· | ···················· |
| Y | 2813 hCoV-19/Belgium/rega-14121333/2020\|EPI_ISL_738386\| | ···················· | ···················· | ····················· | ···················· | ···················· | ···················· |
| Y | 2814 hCoV-19/Belgium/rega-12141334/2020\|EPI_ISL_738387\| | ···················· | ···················· | ····················· | ···················· | ···················· | ···················· |
| Y | 2815 hCoV-19/Belgium/rega-12141335/2020\|EPI_ISL_738388\| | ···················· | ···················· | ····················· | ···················· | ···················· | ···················· |
| Y | 2816 hCoV-19/Belgium/rega-12091336/2020\|EPI_ISL_738389\| | ···················· | ···················· | ····················· | ···················· | ···················· | ···················· |
| Y | 2817 hCoV-19/England/NORW-21FF4B/2020\|EPI_ISL_725080\|20 | ···················· | ···················· | ····················· | ···················· | ···················· | ···················· |
| Y | 2818 hCoV-19/England/NORW-220C40/2020\|EPI_ISL_725082\|20 | ···················· | ·A·················· | ····················· | ···················· | ···················· | ···A················ |
| Y | 2819 hCoV-19/England/NORW-220CE6/2020\|EPI_ISL_725085\|20 | ···················· | ·A·················· | ····················· | ···················· | ···················· | ···A················ |
| Y | 2820 hCoV-19/England/NORW-220D2F/2020\|EPI_ISL_725088\|20 | ···················· | ···················· | ····················· | ···················· | ···················· | ···················· |
| Y | 2821 hCoV-19/England/NORW-220DB6/2020\|EPI_ISL_725093\|20 | ···················· | ···················· | ····················· | ···················· | ···················· | ···················· |
| Y | 2822 hCoV-19/England/NORW-220DE3/2020\|EPI_ISL_725095\|20 | ···················· | ···················· | ····················· | ···················· | ···················· | ···················· |
| Y | 2823 hCoV-19/England/NORW-220DF2/2020\|EPI_ISL_725096\|20 | ···················· | ···················· | ····················· | ···················· | ···················· | ···················· |
| Y | 2824 hCoV-19/England/NORW-220E3B/2020\|EPI_ISL_725098\|20 | ···················· | ·R·················· | ····················· | ···················· | ···················· | ···R················ |
| Y | 2826 hCoV-19/Portugal/PT1978/2020\|EPI_ISL_732201\|2020-1 | ···················· | ···················· | ····················· | ···················· | ···················· | ···················· |
| Y | 2827 hCoV-19/Portugal/PT1979/2020\|EPI_ISL_732202\|2020-1 | ···················· | ···················· | ····················· | ···················· | ···················· | ···················· |
| Y | 2828 hCoV-19/Portugal/PT1980/2020\|EPI_ISL_732203\|2020-1 | ···················· | ···················· | ····················· | ···················· | ···················· | ···················· |
| Y | 2829 hCoV-19/Portugal/PT1981/2020\|EPI_ISL_732204\|2020-1 | ···················· | ···················· | ····················· | ···················· | ···················· | ···················· |
| Y | 2830 hCoV-19/Portugal/PT1982/2020\|EPI_ISL_732205\|2020-1 | ···················· | ···················· | ····················· | ···················· | ···················· | ···················· |
| Y | 2831 hCoV-19/Portugal/PT1983/2020\|EPI_ISL_732206\|2020-1 | ···················· | ···················· | ····················· | ···················· | ···················· | ···················· |
| Y | 2832 hCoV-19/Portugal/PT1984/2020\|EPI_ISL_732207\|2020-1 | ···················· | ···················· | ····················· | ···················· | ···················· | ···················· |
| Y | 2833 hCoV-19/Portugal/PT1985/2020\|EPI_ISL_732208\|2020-1 | ···················· | ···················· | ····················· | ···················· | ···················· | ···················· |
| Y | 2834 hCoV-19/Portugal/PT1986/2020\|EPI_ISL_732209\|2020-1 | ···················· | ···················· | ····················· | ···················· | ···················· | ···················· |
| Y | 2835 hCoV-19/Portugal/PT1987/2020\|EPI_ISL_732210\|2020-1 | ···················· | ···················· | ····················· | ···················· | ···················· | ···················· |
| Y | 2836 hCoV-19/Portugal/PT1988/2020\|EPI_ISL_732211\|2020-1 | ···················· | ···················· | ····················· | ···················· | ···················· | ···················· |
| Y | 2837 hCoV-19/Portugal/PT1990/2020\|EPI_ISL_732212\|2020-1 | ···················· | ···················· | ····················· | ···················· | ···················· | ···················· |
| Y | 2838 hCoV-19/Portugal/PT1991/2020\|EPI_ISL_732213\|2020-1 | ···················· | ···················· | ····················· | ···················· | ···················· | ···················· |
| Y | 2839 hCoV-19/Portugal/PT1992/2020\|EPI_ISL_732214\|2020-1 | ···················· | ···················· | ····················· | ···················· | ···················· | ···················· |
| Y | 2840 hCoV-19/Portugal/PT1993/2020\|EPI_ISL_732215\|2020-1 | ···················· | ···················· | ····················· | ···················· | ···················· | ···················· |
| Y | 2841 hCoV-19/Portugal/PT1995/2020\|EPI_ISL_732216\|2020-1 | ···················· | ···················· | ····················· | ···················· | ···················· | ···················· |
| Y | 2842 hCoV-19/Portugal/PT1996/2020\|EPI_ISL_732217\|2020-1 | ···················· | ···················· | ····················· | ···················· | ···················· | ···················· |
| Y | 2843 hCoV-19/Portugal/PT1997/2020\|EPI_ISL_732218\|2020-1 | ···················· | ···················· | ····················· | ···················· | ···················· | ···················· |
| Y | 2844 hCoV-19/Portugal/PT1998/2020\|EPI_ISL_732219\|2020-1 | ···················· | ···················· | ····················· | ···················· | ···················· | ···················· |
| Y | 2845 hCoV-19/Portugal/PT1999/2020\|EPI_ISL_732220\|2020-1 | ···················· | ···················· | ····················· | ···················· | ···················· | ···················· |
| Y | 2846 hCoV-19/Portugal/PT2000/2020\|EPI_ISL_732221\|2020-1 | ···················· | ···················· | ····················· | ···················· | ···················· | ···················· |
| Y | 2847 hCoV-19/Portugal/PT2001/2020\|EPI_ISL_732222\|2020-1 | ···················· | ···················· | ····················· | ···················· | ···················· | ···················· |
| Y | 2848 hCoV-19/Portugal/PT2002/2020\|EPI_ISL_732223\|2020-1 | ···················· | ···················· | ····················· | ···················· | ···················· | ···················· |
| Y | 2849 hCoV-19/Portugal/PT2003/2020\|EPI_ISL_732224\|2020-1 | ···················· | ···················· | ····················· | ···················· | ···················· | ···················· |
| Y | 2850 hCoV-19/Portugal/PT2004/2020\|EPI_ISL_732225\|2020-1 | ···················· | ···················· | ····················· | ···················· | ···················· | ···················· |
| Y | 2851 hCoV-19/Portugal/PT2005/2020\|EPI_ISL_732226\|2020-1 | ···················· | ···················· | ····················· | ···················· | ···················· | ···················· |
| Y | 2852 hCoV-19/Portugal/PT2006/2020\|EPI_ISL_732227\|2020-1 | ···················· | ···················· | ····················· | ···················· | ···················· | ···················· |
| Y | 2853 hCoV-19/Portugal/PT2007/2020\|EPI_ISL_732228\|2020-1 | ···················· | ···················· | ····················· | ···················· | ···················· | ···················· |
| Y | 2854 hCoV-19/Portugal/PT2008/2020\|EPI_ISL_732229\|2020-1 | ···················· | ···················· | ····················· | ···················· | ···················· | ···················· |
| Y | 2855 hCoV-19/Portugal/PT2009/2020\|EPI_ISL_732230\|2020-1 | ···················· | ···················· | ····················· | ···················· | ···················· | ···················· |
| Y | 2856 hCoV-19/Portugal/PT2010/2020\|EPI_ISL_732231\|2020-1 | ···················· | ···················· | ····················· | ···················· | ···················· | ···················· |
| Y | 2857 hCoV-19/Portugal/PT2011/2020\|EPI_ISL_732232\|2020-1 | ···················· | ···················· | ····················· | ···················· | ···················· | ···················· |
| Y | 2858 hCoV-19/Portugal/PT2014/2020\|EPI_ISL_732233\|2020-1 | ···················· | ···················· | ····················· | ···················· | ···················· | ···················· |
| Y | 2859 hCoV-19/Portugal/PT2015/2020\|EPI_ISL_732234\|2020-1 | ···················· | ···················· | ····················· | ···················· | ···················· | ···················· |
| Y | 2860 hCoV-19/Portugal/PT2016/2020\|EPI_ISL_732235\|2020-1 | ···················· | ···················· | ····················· | ···················· | ···················· | ···················· |
| Y | 2861 hCoV-19/Portugal/PT2017/2020\|EPI_ISL_732236\|2020-1 | ···················· | ···················· | ····················· | ···················· | ···················· | ···················· |
| Y | 2862 hCoV-19/Portugal/PT2018/2020\|EPI_ISL_732237\|2020-1 | ···················· | ···················· | ····················· | ···················· | ···················· | ···················· |
| Y | 2863 hCoV-19/Portugal/PT2019/2020\|EPI_ISL_732238\|2020-1 | ···················· | ···················· | ····················· | ···················· | ···················· | ···················· |
| Y | 2864 hCoV-19/Portugal/PT2020/2020\|EPI_ISL_732239\|2020-1 | ···················· | ···················· | ····················· | ···················· | ···················· | ···················· |
| Y | 2865 hCoV-19/Portugal/PT2021/2020\|EPI_ISL_732240\|2020-1 | ···················· | ···················· | ····················· | ···················· | ···················· | ···················· |
| Y | 2866 hCoV-19/Portugal/PT2023/2020\|EPI_ISL_732241\|2020-1 | ···················· | ···················· | ····················· | ···················· | ···················· | ···················· |
| Y | 2867 hCoV-19/Portugal/PT2024/2020\|EPI_ISL_732242\|2020-1 | ···················· | ···················· | ····················· | ···················· | ···················· | ···················· |
| Y | 2868 hCoV-19/Portugal/PT2027/2020\|EPI_ISL_732243\|2020-1 | ···················· | ···················· | ····················· | ···················· | ···················· | ···················· |
| Y | 2869 hCoV-19/Portugal/PT2028/2020\|EPI_ISL_732244\|2020-1 | ···················· | ···················· | ····················· | ···················· | ···················· | ···················· |
| Y | 2870 hCoV-19/Portugal/PT2029/2020\|EPI_ISL_732245\|2020-1 | ···················· | ···················· | ····················· | ···················· | ···················· | ···················· |
| Y | 2871 hCoV-19/Portugal/PT2030/2020\|EPI_ISL_732246\|2020-1 | ···················· | ·········A·········· | ····················· | ···················· | ······T············· | ···················· |
| Y | 2872 hCoV-19/Portugal/PT2031/2020\|EPI_ISL_732247\|2020-1 | ···················· | ···················· | ····················· | ···················· | ···················· | ···················· |
| Y | 2873 hCoV-19/Portugal/PT2032/2020\|EPI_ISL_732248\|2020-1 | ···················· | ···················· | ····················· | ···················· | ···················· | ···················· |
| Y | 2874 hCoV-19/Portugal/PT2033/2020\|EPI_ISL_732249\|2020-1 | ···················· | ···················· | ····················· | ···················· | ···················· | ···················· |

Y 2876 hCoV-19/Portugal/PT2064/2020|EPI_ISL_732278|2020-1 ···················· ···················· ····················· ···················· ···················· ····················
Y 2877 hCoV-19/Portugal/PT2066/2020|EPI_ISL_732280|2020-1 ···················· ···················· ····················· ···················· ···················· ····················
Y 2878 hCoV-19/Portugal/PT2067/2020|EPI_ISL_732281|2020-1 ···················· ···················· ····················· ···················· ···················· ····················
Y 2879 hCoV-19/Portugal/PT2068/2020|EPI_ISL_732282|2020-1 ···················· ···················· ····················· ···················· ···················· ····················
Y 2880 hCoV-19/Portugal/PT2069/2020|EPI_ISL_732283|2020-1 ···················· ···················· ····················· ···················· ···················· ····················
Y 2881 hCoV-19/Portugal/PT2070/2020|EPI_ISL_732284|2020-1 ···················· ···················· ····················· ···················· ···················· ····················
Y 2882 hCoV-19/Portugal/PT2071/2020|EPI_ISL_732285|2020-1 ···················· ···················· ····················· ···················· ···················· ····················
Y 2883 hCoV-19/Portugal/PT2072/2020|EPI_ISL_732286|2020-1 ···················· ···················· ····················· ···················· ···················· ····················
Y 2884 hCoV-19/Portugal/PT2073/2020|EPI_ISL_732287|2020-1 ···················· ···················· ····················· ···················· ···················· ····················
Y 2885 hCoV-19/Portugal/PT2074/2020|EPI_ISL_732288|2020-1 ···················· ···················· ····················· ···················· ···················· ····················
Y 2886 hCoV-19/Portugal/PT2075/2020|EPI_ISL_732289|2020-1 ···················· ···················· ····················· ···················· ···················· ····················
Y 2887 hCoV-19/Tunisia/19695/2020|EPI_ISL_733500|2020-07- ···················· ···················· ····················· ···················· ···················· ····················
Y 2888 hCoV-19/Germany/NW-HHU-439/2020|EPI_ISL_733502|202 ···················· ···················· ····················· ···················· ···················· ····················
Y 2889 hCoV-19/Germany/NW-HHU-447/2020|EPI_ISL_733503|202 ···················· ···················· ····················· ···················· ···················· ····················
Y 2890 hCoV-19/Germany/NW-HHU-448/2020|EPI_ISL_733504|202 ···················· ·····A·············· ····················· ···················· ···················· ····················
Y 2891 hCoV-19/Germany/NW-HHU-451/2020|EPI_ISL_733505|202 ···················· ···················· ····················· ···················· ···················· ····················
Y 2892 hCoV-19/Germany/NW-HHU-431/2020|EPI_ISL_733506|202 ···················· ···················· ····················· ···················· ···················· ····················
Y 2893 hCoV-19/Germany/NW-HHU-432/2020|EPI_ISL_733507|202 ···················· ···················· ····················· ···················· ···················· ····················
Y 2894 hCoV-19/Germany/NW-HHU-433/2020|EPI_ISL_733508|202 ···················· ···················· ····················· ···················· ···················· ····················
Y 2895 hCoV-19/Germany/NW-HHU-434/2020|EPI_ISL_733509|202 ···················· ···················· ····················· ···················· ···················· ····················
Y 2896 hCoV-19/Germany/NW-HHU-435/2020|EPI_ISL_733510|202 ···················· ···················· ····················· ···················· ···················· ····················
Y 2897 hCoV-19/Germany/NW-HHU-436/2020|EPI_ISL_733511|202 ···················· ···················· ····················· ···················· ···················· ····················
Y 2898 hCoV-19/Germany/NW-HHU-437/2020|EPI_ISL_733512|202 ···················· ···················· ····················· ···················· ···················· ····················
Y 2899 hCoV-19/Germany/NW-HHU-438/2020|EPI_ISL_733513|202 ···················· ···················· ····················· ···················· ···················· ····················
Y 2900 hCoV-19/Germany/NW-HHU-440/2020|EPI_ISL_733514|202 ···················· ···················· ····················· ···················· ···················· ····················
Y 2901 hCoV-19/Germany/NW-HHU-442/2020|EPI_ISL_733515|202 ···················· ···················· ····················· ···················· ···················· ····················
Y 2902 hCoV-19/Germany/NW-HHU-443/2020|EPI_ISL_733516|202 ···················· ···················· ····················· ···················· ···················· ····················
Y 2903 hCoV-19/Germany/NW-HHU-444/2020|EPI_ISL_733517|202 ···················· ···················· ····················· ···················· ···················· ····················
Y 2904 hCoV-19/Germany/NW-HHU-445/2020|EPI_ISL_733518|202 ···················· ···················· ····················· ···················· ···················· ····················
Y 2905 hCoV-19/Germany/NW-HHU-446/2020|EPI_ISL_733519|202 ···················· ·A·················· ····················· ···················· ···················· ····················
Y 2906 hCoV-19/Germany/NW-HHU-449/2020|EPI_ISL_733520|202 ···················· ···················· ····················· ···················· ···················· ····················
Y 2907 hCoV-19/Germany/NW-HHU-450/2020|EPI_ISL_733521|202 ···················· ···················· ····················· ···················· ···················· ····················
Y 2908 hCoV-19/Germany/NW-HHU-452/2020|EPI_ISL_733522|202 ···················· ···················· ····················· ···················· ···················· ····················
Y 2909 hCoV-19/Germany/NW-HHU-453/2020|EPI_ISL_733523|202 ···················· ···················· ····················· ···················· ···················· ····················
Y 2910 hCoV-19/Germany/NW-HHU-317/2020|EPI_ISL_733525|202 ···················· ···················· ····················· ···················· ···················· ····················
Y 2911 hCoV-19/Germany/NW-HHU-318/2020|EPI_ISL_733526|202 ···················· ·A·················· ····················· ···················· ···················· ··A·················
Y 2912 hCoV-19/Germany/NW-HHU-319/2020|EPI_ISL_733527|202 ···················· ·A·················· ····················· ···················· ···················· ··A·················
Y 2948 hCoV-19/env/Hong Kong/VN20001008/2020|EPI_ISL_7335 ···················· ·A·················· ····················· ···················· ···················· ··A·················
Y 2949 hCoV-19/Hong Kong/VB20275039/2020|EPI_ISL_733569|2 ···················· ·A·················· ····················· ···················· ···················· ··A·················
Y 2950 hCoV-19/Hong Kong/VM20192019/2020|EPI_ISL_733570|2 ···················· ·A·················· ····················· ···················· ···················· ··A·················
Y 2951 hCoV-19/Hong Kong/VM20149166/2020|EPI_ISL_733571|2 ···················· ···················· ····················· ···················· ···················· ····················
Y 2952 hCoV-19/Hong Kong/CM20000444/2020|EPI_ISL_733572|2 ···················· ···················· ····················· ···················· ···················· ····················
Y 2953 hCoV-19/Hong Kong/CM20000221/2020|EPI_ISL_733574|2 ···················· ···················· ····················· ···················· ···················· ····················
Y 2954 hCoV-19/Hong Kong/CM20000417/2020|EPI_ISL_733575|2 ···················· ···················· ····················· ···················· ···················· ····················
Y 2955 hCoV-19/Hong Kong/CM20000368/2020|EPI_ISL_733576|2 ···················· ···················· ····················· ···················· ···················· ····················
Y 2956 hCoV-19/Hong Kong/CM20000418/2020|EPI_ISL_733579|2 ···················· ···················· ····················· ···················· ···················· ····················
Y 2957 hCoV-19/Belgium/rega-0717849/2020|EPI_ISL_734800|2 ···················· ···················· ····················· ···················· ···················· ····················
Y 2958 hCoV-19/Belgium/rega-0719696/2020|EPI_ISL_734801|2 ···················· ···················· ····················· ···················· ···················· ····················
Y 2959 hCoV-19/Belgium/rega-0721811/2020|EPI_ISL_734804|2 ···················· ···················· ····················· ···················· ···················· ····················
Y 2960 hCoV-19/Belgium/rega-0721812/2020|EPI_ISL_734805|2 ···················· ···················· ····················· ···················· ···················· ····················
Y 2961 hCoV-19/Belgium/rega-0721840/2020|EPI_ISL_734810|2 ···················· ···················· ····················· ···················· ···················· ····················
Y 2962 hCoV-19/Belgium/rega-0721841/2020|EPI_ISL_734811|2 ···················· ···················· ····················· ···················· ···················· ····················
Y 2963 hCoV-19/Belgium/rega-0721842/2020|EPI_ISL_734812|2 ···················· ···················· ····················· ···················· ···················· ····················
Y 2964 hCoV-19/Belgium/rega-0722829/2020|EPI_ISL_734824|2 ···················· ···················· ····················· ···················· ···················· ····················
Y 2965 hCoV-19/Belgium/rega-0723820/2020|EPI_ISL_734825|2 ···················· ···················· ····················· ···················· ···················· ····················
Y 2966 hCoV-19/Belgium/rega-0723821/2020|EPI_ISL_734826|2 ···················· ···················· ····················· ···················· ···················· ····················
Y 2967 hCoV-19/Belgium/rega-0729922/2020|EPI_ISL_734827|2 ···················· ···················· ····················· ···················· ···················· ····················
Y 2968 hCoV-19/Belgium/rega-0731777/2020|EPI_ISL_734828|2 ···················· ···················· ····················· ···················· ···················· ····················
Y 2969 hCoV-19/Belgium/rega-0731778/2020|EPI_ISL_734829|2 ···················· ···················· ····················· ···················· ···················· ····················
Y 2970 hCoV-19/Belgium/rega-0731779/2020|EPI_ISL_734830|2 ···················· ···················· ····················· ···················· ···················· ····················
Y 2971 hCoV-19/Belgium/rega-0731780/2020|EPI_ISL_734831|2 ···················· ···················· ····················· ···················· ···················· ····················
Y 2972 hCoV-19/Belgium/rega-0731781/2020|EPI_ISL_734832|2 ···················· ···················· ····················· ···················· ···················· ····················
Y 2973 hCoV-19/Belgium/rega-0731782/2020|EPI_ISL_734833|2 ···················· ···················· ····················· ···················· ···················· ····················
Y 2974 hCoV-19/Belgium/rega-0731785/2020|EPI_ISL_734834|2 ···················· ···················· ····················· ···················· ···················· ····················
Y 2975 hCoV-19/Belgium/rega-0731787/2020|EPI_ISL_734835|2 ···················· ···················· ····················· ···················· ···················· ····················
Y 2976 hCoV-19/Belgium/rega-0731789/2020|EPI_ISL_734836|2 ···················· ···················· ····················· ···················· ···················· ····················
Y 2977 hCoV-19/Belgium/rega-0731790/2020|EPI_ISL_734837|2 ···················· ···················· ····················· ···················· ···················· ····················
Y 2978 hCoV-19/Belgium/rega-0731791/2020|EPI_ISL_734838|2 ···················· ···················· ····················· ···················· ···················· ····················
Y 2979 hCoV-19/Belgium/rega-0731792/2020|EPI_ISL_734839|2 ···················· ···················· ····················· ···················· ···················· ····················
Y 2980 hCoV-19/Belgium/rega-0801752/2020|EPI_ISL_734840|2 ···················· ···················· ····················· ···················· ···················· ····················
Y 2981 hCoV-19/Belgium/rega-0801754/2020|EPI_ISL_734841|2 ···················· ···················· ····················· ···················· ···················· ····················
Y 2982 hCoV-19/Belgium/rega-0801761/2020|EPI_ISL_734842|2 ···················· ···················· ····················· ···················· ···················· ····················
Y 2983 hCoV-19/Belgium/rega-0801762/2020|EPI_ISL_734843|2 ···················· ···················· ····················· ···················· ···················· ····················
Y 2984 hCoV-19/Belgium/rega-0801767/2020|EPI_ISL_734844|2 ···················· ···················· ····················· ···················· ···················· ····················
Y 2985 hCoV-19/Belgium/rega-0801768/2020|EPI_ISL_734845|2 ···················· ···················· ····················· ···················· ···················· ····················
Y 2986 hCoV-19/Belgium/rega-0801769/2020|EPI_ISL_734846|2 ···················· ···················· ····················· ···················· ···················· ····················
Y 2987 hCoV-19/Belgium/rega-0801770/2020|EPI_ISL_734847|2 ···················· ···················· ····················· ···················· ···················· ····················
Y 2988 hCoV-19/Belgium/rega-0801771/2020|EPI_ISL_734848|2 ···················· ···················· ····················· ···················· ···················· ····················
Y 2989 hCoV-19/Belgium/rega-0801772/2020|EPI_ISL_734849|2 ···················· ···················· ····················· ···················· ···················· ····················
Y 2990 hCoV-19/Belgium/rega-0801773/2020|EPI_ISL_734850|2 ···················· ···················· ····················· ···················· ···················· ····················

| | | | | | | | | |
|---|---|---|---|---|---|---|---|---|
| Y | 2991 | hCoV-19/Belgium/rega-0801774/2020\|EPI_ISL_734851\|2 | ···················· | ···················· | ····················· | ···················· | ···················· | ···················· |
| Y | 2992 | hCoV-19/Belgium/rega-0801783/2020\|EPI_ISL_734852\|2 | ···················· | ···················· | ····················· | ···················· | ···················· | ···················· |
| Y | 2993 | hCoV-19/Belgium/rega-0801784/2020\|EPI_ISL_734853\|2 | ···················· | ···················· | ····················· | ···················· | ···················· | ···················· |
| Y | 2994 | hCoV-19/Belgium/rega-0801786/2020\|EPI_ISL_734854\|2 | ···················· | ···················· | ····················· | ···················· | ···················· | ···················· |
| Y | 2995 | hCoV-19/Belgium/rega-0801788/2020\|EPI_ISL_734855\|2 | ···················· | ···················· | ····················· | ···················· | ···················· | ···················· |
| Y | 2996 | hCoV-19/Belgium/rega-0801793/2020\|EPI_ISL_734856\|2 | ···················· | ···················· | ····················· | ···················· | ···················· | ···················· |
| Y | 2997 | hCoV-19/Belgium/rega-0801794/2020\|EPI_ISL_734857\|2 | ···················· | ···················· | ····················· | ···················· | ···················· | ···················· |
| Y | 2998 | hCoV-19/Belgium/rega-0801795/2020\|EPI_ISL_734858\|2 | ···················· | ···················· | ····················· | ···················· | ···················· | ···················· |
| Y | 2999 | hCoV-19/Belgium/rega-0801796/2020\|EPI_ISL_734859\|2 | ···················· | ···················· | ····················· | ···················· | ···················· | ···················· |
| Y | 3000 | hCoV-19/Belgium/rega-0801797/2020\|EPI_ISL_734860\|2 | ···················· | ···················· | ····················· | ···················· | ···················· | ···················· |
| Y | 3001 | hCoV-19/Belgium/rega-0801798/2020\|EPI_ISL_734861\|2 | ···················· | ···················· | ····················· | ···················· | ···················· | ···················· |
| Y | 3002 | hCoV-19/Belgium/rega-0801799/2020\|EPI_ISL_734862\|2 | ···················· | ···················· | ····················· | ···················· | ···················· | ···················· |
| Y | 3003 | hCoV-19/Belgium/rega-0801800/2020\|EPI_ISL_734863\|2 | ···················· | ···················· | ····················· | ···················· | ···················· | ···················· |
| Y | 3004 | hCoV-19/Belgium/rega-0801801/2020\|EPI_ISL_734864\|2 | ···················· | ···················· | ····················· | ···················· | ···················· | ···················· |
| Y | 3005 | hCoV-19/Belgium/rega-0801923/2020\|EPI_ISL_734865\|2 | ···················· | ···················· | ····················· | ···················· | ···················· | ···················· |
| Y | 3006 | hCoV-19/Belgium/rega-0801924/2020\|EPI_ISL_734866\|2 | ···················· | ···················· | ····················· | ···················· | ···················· | ···················· |
| Y | 3007 | hCoV-19/Belgium/rega-0801925/2020\|EPI_ISL_734867\|2 | ···················· | ···················· | ····················· | ···················· | ···················· | ···················· |
| Y | 3008 | hCoV-19/Belgium/rega-0801926/2020\|EPI_ISL_734868\|2 | ···················· | ···················· | ····················· | ···················· | ···················· | ···················· |
| Y | 3009 | hCoV-19/Belgium/rega-0801927/2020\|EPI_ISL_734869\|2 | ···················· | ···················· | ····················· | ···················· | ···················· | ···················· |
| Y | 3010 | hCoV-19/Belgium/rega-0801928/2020\|EPI_ISL_734870\|2 | ···················· | ···················· | ····················· | ···················· | ···················· | ···················· |
| Y | 3011 | hCoV-19/Belgium/rega-0802749/2020\|EPI_ISL_734871\|2 | ···················· | ···················· | ····················· | ···················· | ···················· | ···················· |
| Y | 3012 | hCoV-19/Belgium/rega-0802750/2020\|EPI_ISL_734872\|2 | ···················· | ···················· | ····················· | ···················· | ···················· | ···················· |
| Y | 3013 | hCoV-19/Belgium/rega-0802763/2020\|EPI_ISL_734873\|2 | ···················· | ···················· | ····················· | ···················· | ···················· | ···················· |
| Y | 3014 | hCoV-19/Belgium/rega-0802764/2020\|EPI_ISL_734874\|2 | ···················· | ···················· | ····················· | ···················· | ···················· | ···················· |
| Y | 3015 | hCoV-19/Belgium/rega-0802775/2020\|EPI_ISL_734875\|2 | ···················· | ···················· | ····················· | ···················· | ···················· | ···················· |
| Y | 3016 | hCoV-19/Belgium/rega-0802776/2020\|EPI_ISL_734876\|2 | ···················· | ···················· | ····················· | ···················· | ···················· | ···················· |
| Y | 3017 | hCoV-19/Belgium/rega-0803720/2020\|EPI_ISL_734877\|2 | ···················· | ···················· | ····················· | ···················· | ···················· | ···················· |
| Y | 3018 | hCoV-19/Belgium/rega-0803721/2020\|EPI_ISL_734878\|2 | ···················· | ···················· | ····················· | ···················· | ···················· | ···················· |
| Y | 3019 | hCoV-19/Belgium/rega-0803722/2020\|EPI_ISL_734879\|2 | ···················· | ···················· | ····················· | ···················· | ···················· | ···················· |
| Y | 3020 | hCoV-19/Belgium/rega-0803723/2020\|EPI_ISL_734880\|2 | ···················· | ···················· | ····················· | ···················· | ···················· | ···················· |
| Y | 3021 | hCoV-19/Belgium/rega-0803724/2020\|EPI_ISL_734881\|2 | ···················· | ···················· | ····················· | ···················· | ···················· | ···················· |
| Y | 3022 | hCoV-19/Belgium/rega-0803725/2020\|EPI_ISL_734882\|2 | ···················· | ···················· | ····················· | ···················· | ···················· | ···················· |
| Y | 3023 | hCoV-19/Belgium/rega-0803727/2020\|EPI_ISL_734883\|2 | ···················· | ···················· | ····················· | ···················· | ···················· | ···················· |
| Y | 3024 | hCoV-19/Belgium/rega-0803730/2020\|EPI_ISL_734884\|2 | ···················· | ···················· | ····················· | ···················· | ···················· | ···················· |
| Y | 3025 | hCoV-19/Belgium/rega-0803732/2020\|EPI_ISL_734885\|2 | ···················· | ···················· | ····················· | ···················· | ···················· | ···················· |
| Y | 3026 | hCoV-19/Belgium/rega-0803733/2020\|EPI_ISL_734886\|2 | ···················· | ···················· | ····················· | ···················· | ···················· | ···················· |
| Y | 3027 | hCoV-19/Belgium/rega-0803734/2020\|EPI_ISL_734887\|2 | ···················· | ···················· | ····················· | ···················· | ···················· | ···················· |
| Y | 3028 | hCoV-19/Belgium/rega-0803736/2020\|EPI_ISL_734888\|2 | ···················· | ····A··············· | ····················· | ···················· | ···················· | ···················· |
| Y | 3029 | hCoV-19/Belgium/rega-0803737/2020\|EPI_ISL_734889\|2 | ···················· | ···················· | ····················· | ···················· | ···················· | ···················· |
| Y | 3030 | hCoV-19/Belgium/rega-0803739/2020\|EPI_ISL_734890\|2 | ···················· | ···················· | ····················· | ···················· | ···················· | ···················· |
| Y | 3031 | hCoV-19/Belgium/rega-0803740/2020\|EPI_ISL_734891\|2 | ···················· | ···················· | ····················· | ···················· | ···················· | ···················· |
| Y | 3032 | hCoV-19/Belgium/rega-0803741/2020\|EPI_ISL_734892\|2 | ···················· | ···················· | ····················· | ···················· | ···················· | ···················· |
| Y | 3033 | hCoV-19/Belgium/rega-0803745/2020\|EPI_ISL_734893\|2 | ···················· | ···················· | ····················· | ···················· | ···················· | ···················· |
| Y | 3034 | hCoV-19/Belgium/rega-0803746/2020\|EPI_ISL_734894\|2 | ···················· | ···················· | ····················· | ···················· | ···················· | ···················· |
| Y | 3035 | hCoV-19/Belgium/rega-0803747/2020\|EPI_ISL_734895\|2 | ···················· | ···················· | ····················· | ···················· | ···················· | ···················· |
| Y | 3036 | hCoV-19/Belgium/rega-0803751/2020\|EPI_ISL_734896\|2 | ···················· | ···················· | ····················· | ···················· | ···················· | ···················· |
| Y | 3037 | hCoV-19/Belgium/rega-0803753/2020\|EPI_ISL_734897\|2 | ···················· | ···················· | ····················· | ···················· | ···················· | ···················· |
| Y | 3038 | hCoV-19/Belgium/rega-0803755/2020\|EPI_ISL_734898\|2 | ···················· | ···················· | ····················· | ···················· | ···················· | ···················· |
| Y | 3039 | hCoV-19/Belgium/rega-0803756/2020\|EPI_ISL_734899\|2 | ···················· | ···················· | ····················· | ···················· | ···················· | ···················· |
| Y | 3040 | hCoV-19/Finland/S9BC1/2020\|EPI_ISL_737060\|2020-10- | ···················· | ···················· | ····················· | ···················· | ···················· | ···················· |
| Y | 3041 | hCoV-19/Finland/S12A1/2020\|EPI_ISL_737061\|2020-09- | ···················· | ···················· | ····················· | ···················· | ···················· | ···················· |
| Y | 3042 | hCoV-19/Finland/S12F7/2020\|EPI_ISL_737062\|2020-09- | ···················· | ···················· | ····················· | ···················· | ···················· | ···················· |
| Y | 3043 | hCoV-19/Finland/S20B7/2020\|EPI_ISL_737063\|2020-09- | ···················· | ·A·················· | ····················· | ···················· | ···················· | ···A················ |
| Y | 3044 | hCoV-19/Finland/S5C4/2020\|EPI_ISL_737064\|2020-09-1 | ···················· | ···················· | ····················· | ···················· | ···················· | ···················· |
| Y | 3045 | hCoV-19/Finland/S5D6/2020\|EPI_ISL_737065\|2020-09-1 | ···················· | ···················· | ····················· | ···················· | ···················· | ···················· |
| Y | 3046 | hCoV-19/Finland/S5E7/2020\|EPI_ISL_737066\|2020-09-1 | ···················· | ···················· | ····················· | ···················· | ···················· | ···················· |
| Y | 3047 | hCoV-19/Finland/S5F8/2020\|EPI_ISL_737067\|2020-09-1 | ···················· | ···················· | ····················· | ···················· | ···················· | ···················· |
| Y | 3048 | hCoV-19/Finland/S5G2/2020\|EPI_ISL_737068\|2020-09-1 | ···················· | ···················· | ····················· | ···················· | ···················· | ···················· |
| Y | 3049 | hCoV-19/Finland/S14G1/2020\|EPI_ISL_737069\|2020-08- | ···················· | ···················· | ····················· | ···················· | ···················· | ···················· |
| Y | 3050 | hCoV-19/Finland/S11AA3/2020\|EPI_ISL_737070\|2020-10 | ···················· | ·A·················· | ····················· | ···················· | ···················· | ···A················ |
| Y | 3051 | hCoV-19/Finland/S11AF5/2020\|EPI_ISL_737071\|2020-10 | ···················· | ···················· | ····················· | ···················· | ···················· | ···················· |
| Y | 3052 | hCoV-19/Finland/S12H7/2020\|EPI_ISL_737072\|2020-10- | ···················· | ·A·················· | ····················· | ···················· | ···················· | ···A················ |
| Y | 3053 | hCoV-19/Finland/S16BC7/2020\|EPI_ISL_737073\|2020-10 | ···················· | ···················· | ····················· | ···················· | ···················· | ···················· |
| Y | 3054 | hCoV-19/Finland/S17AD11/2020\|EPI_ISL_737074\|2020-1 | ···················· | ·A·················· | ····················· | ···················· | ···················· | ···A················ |
| Y | 3055 | hCoV-19/Finland/S19AB3/2020\|EPI_ISL_737075\|2020-10 | ···················· | ···················· | ····················· | ···················· | ···················· | ···················· |
| Y | 3056 | hCoV-19/Finland/S20AE7/2020\|EPI_ISL_737076\|2020-10 | ···················· | ···················· | ····················· | ···················· | ···················· | ···················· |
| Y | 3057 | hCoV-19/Finland/S22AB11/2020\|EPI_ISL_737077\|2020-1 | ···················· | ···················· | ····················· | ···················· | ···················· | ···················· |
| Y | 3058 | hCoV-19/Finland/S22AB1/2020\|EPI_ISL_737078\|2020-10 | ···················· | ·A·················· | ····················· | ···················· | ···················· | ···A················ |
| Y | 3059 | hCoV-19/Finland/S6AD1/2020\|EPI_ISL_737079\|2020-10- | ···················· | ·A·················· | ····················· | ···················· | ···················· | ···A················ |
| Y | 3060 | hCoV-19/USA/WA-UW-32754/2020\|EPI_ISL_737087\|2020-1 | ···················· | ·A·················· | ····················· | ···················· | ···················· | ···················· |
| Y | 3061 | hCoV-19/USA/WA-UW-32807/2020\|EPI_ISL_737088\|2020-1 | ···················· | ·A·················· | ····················· | ···················· | ···················· | ···················· |
| Y | 3062 | hCoV-19/USA/WA-UW-32819/2020\|EPI_ISL_737089\|2020-1 | ···················· | ·A·················· | ····················· | ···················· | ···················· | ···················· |
| Y | 3063 | hCoV-19/Belgium/rega-12111337/2020\|EPI_ISL_738390\| | ···················· | ···················· | ····················· | ···················· | ···················· | ···················· |
| Y | 3064 | hCoV-19/Belgium/rega-12111338/2020\|EPI_ISL_738391\| | ···················· | ···················· | ····················· | ···················· | ···················· | ···················· |
| Y | 3065 | hCoV-19/Belgium/rega-12141339/2020\|EPI_ISL_738392\| | ···················· | ···················· | ····················· | ···················· | ···················· | ···················· |
| Y | 3066 | hCoV-19/Belgium/rega-12111340/2020\|EPI_ISL_738393\| | ···················· | ···················· | ····················· | ···················· | ···················· | ···················· |
| Y | 3067 | hCoV-19/Belgium/rega-12141341/2020\|EPI_ISL_738394\| | ···················· | ···················· | ····················· | ···················· | ···················· | ···················· |
| Y | 3068 | hCoV-19/Belgium/rega-12111342/2020\|EPI_ISL_738395\| | ···················· | ···················· | ····················· | ···················· | ···················· | ···················· |
| Y | 3069 | hCoV-19/Belgium/rega-12111343/2020\|EPI_ISL_738396\| | ···················· | ···················· | ····················· | ···················· | ···················· | ···················· |
| Y | 3070 | hCoV-19/Belgium/rega-12111344/2020\|EPI_ISL_738397\| | ···················· | ···················· | ····················· | ···················· | ···················· | ···················· |

| | | | | | | | | |
|---|---|---|---|---|---|---|---|---|
| Y | 3071 hCoV-19/Belgium/rega-12141345/2020\|EPI_ISL_738398\| | .................... | .................... | .................... | .................... | .................... | .................... | |
| Y | 3072 hCoV-19/Belgium/rega-12111346/2020\|EPI_ISL_738399\| | .................... | .................... | .................... | .................... | .................... | .................... | |
| Y | 3073 hCoV-19/England/NORW-220EE0/2020\|EPI_ISL_725101\|20 | .................... | .................... | .................... | .................... | .................... | .................... | |
| Y | 3074 hCoV-19/England/NORW-220F47/2020\|EPI_ISL_725103\|20 | .................... | .................... | .................... | .................... | .................... | .................... | |
| Y | 3075 hCoV-19/England/NORW-EFB9D/2020\|EPI_ISL_725107\|202 | .................... | .................... | .................... | .................... | .................... | .................... | |
| Y | 3076 hCoV-19/England/NORW-EFBD9/2020\|EPI_ISL_725108\|202 | .................... | .................... | .................... | .................... | .................... | .................... | |
| Y | 3077 hCoV-19/England/NORW-EFC4F/2020\|EPI_ISL_725110\|202 | .................... | .................... | .................... | .................... | .................... | .................... | |
| Y | 3078 hCoV-19/England/NORW-EFC9A/2020\|EPI_ISL_725111\|202 | .................... | .A.................. | .................... | .................... | .................... | .................... | |
| Y | 3079 hCoV-19/England/NORW-F1846/2020\|EPI_ISL_725113\|202 | .................... | .................... | .................... | .................... | .................... | .................... | |
| Y | 3080 hCoV-19/England/NORW-F1873/2020\|EPI_ISL_725115\|202 | .................... | .................... | .................... | .................... | .................... | .................... | |
| Y | 3081 hCoV-19/England/NORW-F1882/2020\|EPI_ISL_725116\|202 | .................... | .................... | .................... | .................... | .................... | .................... | |
| Y | 3082 hCoV-19/England/NORW-F1891/2020\|EPI_ISL_725117\|202 | .................... | .................... | .................... | .................... | .................... | .................... | |
| Y | 3083 hCoV-19/England/NORW-F18BF/2020\|EPI_ISL_725119\|202 | .................... | .................... | .................... | .................... | .................... | .................... | |
| Y | 3084 hCoV-19/England/NORW-F18CE/2020\|EPI_ISL_725120\|202 | .................... | .................... | .................... | .................... | .................... | .................... | |
| Y | 3085 hCoV-19/England/NORW-F18FB/2020\|EPI_ISL_725122\|202 | .................... | .................... | .................... | .................... | .................... | .................... | |
| Y | 3086 hCoV-19/England/NORW-F1907/2020\|EPI_ISL_725123\|202 | .................... | .................... | .................... | .................... | .................... | .................... | |
| Y | 3087 hCoV-19/England/NORW-F1961/2020\|EPI_ISL_725125\|202 | .................... | .................... | .................... | .................... | .................... | .................... | |
| Y | 3088 hCoV-19/England/NORW-F19CB/2020\|EPI_ISL_725128\|202 | .................... | .................... | .................... | .................... | .................... | .................... | |
| Y | 3089 hCoV-19/England/NORW-F19E9/2020\|EPI_ISL_725129\|202 | .................... | .................... | .................... | .................... | .................... | .................... | |
| Y | 3090 hCoV-19/England/NORW-F1A13/2020\|EPI_ISL_725130\|202 | .................... | .................... | .................... | .................... | .................... | .................... | |
| Y | 3091 hCoV-19/England/NORW-F1A31/2020\|EPI_ISL_725131\|202 | .................... | .................... | .................... | .................... | .................... | .................... | |
| Y | 3092 hCoV-19/England/NORW-F1A5F/2020\|EPI_ISL_725133\|202 | .................... | .................... | .................... | .................... | ......T............. | .................... | |
| Y | 3093 hCoV-19/England/NORW-F1A9B/2020\|EPI_ISL_725134\|202 | .................... | .................... | .................... | .................... | .................... | .................... | |
| Y | 3094 hCoV-19/England/NORW-F1AE6/2020\|EPI_ISL_725137\|202 | .................... | .................... | .................... | .................... | .................... | .................... | |
| Y | 3095 hCoV-19/England/NORW-F37FC/2020\|EPI_ISL_725139\|202 | .................... | .................... | .................... | .................... | .................... | .................... | |
| Y | 3096 hCoV-19/England/NORW-F3817/2020\|EPI_ISL_725140\|202 | .................... | .................... | .................... | .................... | .................... | .................... | |
| Y | 3097 hCoV-19/England/NORW-F3826/2020\|EPI_ISL_725141\|202 | .................... | .................... | .................... | .................... | .................... | .................... | |
| Y | 3098 hCoV-19/England/NORW-F3844/2020\|EPI_ISL_725143\|202 | .................... | .................... | .................... | .................... | .................... | .................... | |
| Y | 3099 hCoV-19/England/NORW-F397E/2020\|EPI_ISL_725145\|202 | .................... | .................... | .................... | .................... | .................... | .................... | |
| Y | 3100 hCoV-19/England/NORW-F398D/2020\|EPI_ISL_725146\|202 | .................... | .................... | .................... | .................... | .................... | .................... | |
| Y | 3101 hCoV-19/England/NORW-F39C9/2020\|EPI_ISL_725147\|202 | .................... | .A.................. | .................... | .................... | .................... | .................... | |
| Y | 3102 hCoV-19/England/NORW-F39F6/2020\|EPI_ISL_725148\|202 | .................... | .................... | .................... | .................... | .................... | .................... | |
| Y | 3103 hCoV-19/England/NORW-F3A20/2020\|EPI_ISL_725150\|202 | .................... | .................... | .................... | .................... | .................... | .................... | |
| Y | 3104 hCoV-19/England/NORW-F3A3F/2020\|EPI_ISL_725151\|202 | .................... | .................... | .................... | .................... | .................... | .................... | |
| Y | 3105 hCoV-19/England/NORW-F3AA8/2020\|EPI_ISL_725155\|202 | .................... | .................... | .................... | .................... | .................... | .................... | |
| Y | 3106 hCoV-19/England/NORW-F3AC6/2020\|EPI_ISL_725156\|202 | .................... | .................... | .................... | .................... | .................... | .................... | |
| Y | 3107 hCoV-19/England/NORW-F400F/2020\|EPI_ISL_725180\|202 | .................... | .................... | .................... | .................... | .................... | .................... | |
| Y | 3108 hCoV-19/England/NORW-F4078/2020\|EPI_ISL_725184\|202 | .................... | .................... | .................... | .................... | .................... | .................... | |
| Y | 3109 hCoV-19/England/NORW-F4096/2020\|EPI_ISL_725186\|202 | .................... | .................... | .................... | .................... | .................... | .................... | |
| Y | 3110 hCoV-19/England/NORW-F40A5/2020\|EPI_ISL_725187\|202 | .................... | .................... | .................... | .................... | .................... | .................... | |
| Y | 3111 hCoV-19/England/NORW-F40B4/2020\|EPI_ISL_725188\|202 | .................... | .................... | .................... | .................... | .................... | .................... | |
| Y | 3112 hCoV-19/England/NORW-F40C3/2020\|EPI_ISL_725189\|202 | .................... | .................... | .................... | .................... | .................... | .................... | |
| Y | 3114 hCoV-19/Wales/PHWC-495804/2020\|EPI_ISL_726401\|2020 | .................... | .................... | .................... | .................... | .................... | .................... | |
| Y | 3115 hCoV-19/Wales/PHWC-495822/2020\|EPI_ISL_726402\|2020 | .................... | .................... | .................... | .................... | .................... | .................... | |
| Y | 3117 hCoV-19/Wales/PHWC-495840/2020\|EPI_ISL_726404\|2020 | .................... | .................... | .................... | .................... | .................... | .................... | |
| Y | 3121 hCoV-19/Wales/PHWC-4958E6/2020\|EPI_ISL_726411\|2020 | .................... | .................... | .................... | .................... | .................... | .................... | |
| Y | 3122 hCoV-19/Wales/PHWC-4958F5/2020\|EPI_ISL_726412\|2020 | .................... | .................... | .................... | .................... | .................... | .................... | |
| Y | 3123 hCoV-19/Wales/PHWC-495901/2020\|EPI_ISL_726413\|2020 | .................... | .................... | .................... | .................... | .................... | .................... | |
| Y | 3125 hCoV-19/Wales/PHWC-49593E/2020\|EPI_ISL_726416\|2020 | .................... | .................... | .................... | .................... | .................... | .................... | |
| Y | 3126 hCoV-19/Wales/PHWC-49594D/2020\|EPI_ISL_726417\|2020 | .................... | .................... | .................... | .................... | .................... | .................... | |
| Y | 3127 hCoV-19/Wales/PHWC-49596B/2020\|EPI_ISL_726418\|2020 | .................... | .................... | .................... | .................... | .................... | .................... | |
| Y | 3130 hCoV-19/Wales/PHWC-495998/2020\|EPI_ISL_726421\|2020 | .................... | .................... | .................... | .................... | .................... | .................... | |
| Y | 3131 hCoV-19/Wales/PHWC-4959B6/2020\|EPI_ISL_726422\|2020 | .................... | .................... | .................... | .................... | .................... | .................... | |
| Y | 3134 hCoV-19/Wales/PHWC-4959E3/2020\|EPI_ISL_726425\|2020 | .................... | .................... | .................... | .................... | .................... | .................... | |
| Y | 3137 hCoV-19/Wales/PHWC-495A3B/2020\|EPI_ISL_726429\|2020 | .................... | .................... | .................... | .................... | .................... | .................... | |
| Y | 3138 hCoV-19/Wales/PHWC-495A95/2020\|EPI_ISL_726433\|2020 | .................... | .................... | .................... | .................... | .................... | .................... | |
| Y | 3140 hCoV-19/Wales/PHWC-495AC2/2020\|EPI_ISL_726436\|2020 | .................... | .................... | .................... | .................... | .................... | .................... | |
| Y | 3141 hCoV-19/Wales/PHWC-495AD1/2020\|EPI_ISL_726437\|2020 | .................... | ......A............. | .................... | .................... | .................... | .................... | |
| Y | 3142 hCoV-19/Wales/PHWC-495AE0/2020\|EPI_ISL_726438\|2020 | .................... | .................... | .................... | .................... | .................... | .................... | |
| Y | 3144 hCoV-19/Wales/PHWC-495B0B/2020\|EPI_ISL_726440\|2020 | .................... | .................... | .................... | .................... | .................... | .................... | |
| Y | 3145 hCoV-19/Wales/PHWC-495B1A/2020\|EPI_ISL_726441\|2020 | .................... | .................... | .................... | .................... | .................... | .................... | |
| Y | 3147 hCoV-19/Wales/PHWC-495B38/2020\|EPI_ISL_726443\|2020 | .................... | .................... | .................... | .................... | .................... | .................... | |
| Y | 3148 hCoV-19/Wales/PHWC-495B47/2020\|EPI_ISL_726444\|2020 | .................... | .................... | .................... | .................... | NNN................. | .................... | 1 |
| Y | 3151 hCoV-19/Wales/PHWC-495B74/2020\|EPI_ISL_726447\|2020 | .................... | .................... | .................... | .................... | .................... | .................... | |
| Y | 3152 hCoV-19/Wales/PHWC-495B83/2020\|EPI_ISL_726448\|2020 | .................... | .................... | .................... | .................... | .................... | .................... | |
| Y | 3153 hCoV-19/Wales/PHWC-495B92/2020\|EPI_ISL_726449\|2020 | .................... | .................... | .................... | .................... | .................... | .................... | |
| Y | 3154 hCoV-19/Wales/PHWC-495BA1/2020\|EPI_ISL_726450\|2020 | .................... | .................... | .................... | .................... | .................... | .................... | |
| Y | 3157 hCoV-19/Wales/PHWC-495BDE/2020\|EPI_ISL_726453\|2020 | .................... | .................... | .................... | .................... | .................... | .................... | |
| Y | 3158 hCoV-19/Wales/PHWC-495BED/2020\|EPI_ISL_726454\|2020 | .................... | .................... | .................... | .................... | .................... | .................... | |
| Y | 3161 hCoV-19/Wales/PHWC-495C17/2020\|EPI_ISL_726457\|2020 | .................... | .................... | .................... | .................... | .................... | .................... | |
| Y | 3163 hCoV-19/Wales/PHWC-495C44/2020\|EPI_ISL_726460\|2020 | .................... | .................... | .................... | .................... | .................... | .................... | |
| Y | 3164 hCoV-19/Wales/PHWC-495C53/2020\|EPI_ISL_726461\|2020 | .................... | .................... | .................... | .................... | .................... | .................... | |
| Y | 3165 hCoV-19/Wales/PHWC-495C62/2020\|EPI_ISL_726462\|2020 | .................... | .................... | .................... | .................... | .................... | .................... | |
| Y | 3166 hCoV-19/Wales/PHWC-495C71/2020\|EPI_ISL_726463\|2020 | .................... | .................... | .................... | .................... | .................... | .................... | |
| Y | 3167 hCoV-19/Wales/PHWC-495C80/2020\|EPI_ISL_726464\|2020 | .................... | .................... | .................... | .................... | .................... | .................... | |
| Y | 3168 hCoV-19/Wales/PHWC-495C9F/2020\|EPI_ISL_726465\|2020 | .................... | ......A............. | .................... | .................... | .................... | .................... | |
| Y | 3169 hCoV-19/Wales/PHWC-495CAE/2020\|EPI_ISL_726466\|2020 | .................... | ......A............. | .................... | .................... | .................... | .................... | |
| Y | 3170 hCoV-19/Wales/PHWC-495CBD/2020\|EPI_ISL_726467\|2020 | .................... | .................... | .................... | .................... | .................... | .................... | |
| Y | 3172 hCoV-19/Wales/PHWC-495CDB/2020\|EPI_ISL_726469\|2020 | .................... | .................... | .................... | .................... | .................... | .................... | |
| Y | 3173 hCoV-19/Wales/PHWC-495CEA/2020\|EPI_ISL_726470\|2020 | .................... | .................... | .................... | .................... | .................... | .................... | |

| | | | | | | | | |
|---|---|---|---|---|---|---|---|---|
| Y | 3174 | hCoV-19/Wales/PHWC-495CF9/2020\|EPI_ISL_726471\|2020 | ···················· | ···················· | ····················· | ···················· | ···················· | ···················· |
| Y | 3175 | hCoV-19/Wales/PHWC-495D05/2020\|EPI_ISL_726472\|2020 | ···················· | ···················· | ····················· | ···················· | ···················· | ···················· |
| Y | 3176 | hCoV-19/Wales/PHWC-495D14/2020\|EPI_ISL_726473\|2020 | ···················· | ···················· | ····················· | ···················· | ···················· | ···················· |
| Y | 3177 | hCoV-19/Wales/PHWC-495D23/2020\|EPI_ISL_726474\|2020 | ···················· | ···················· | ····················· | ···················· | ···················· | ···················· |
| Y | 3178 | hCoV-19/Wales/PHWC-495D32/2020\|EPI_ISL_726475\|2020 | ···················· | ···················· | ····················· | ···················· | ···················· | ···················· |
| Y | 3179 | hCoV-19/Wales/PHWC-495D41/2020\|EPI_ISL_726476\|2020 | ···················· | ···················· | ····················· | ···················· | ···················· | ···················· |
| Y | 3180 | hCoV-19/Wales/PHWC-495D50/2020\|EPI_ISL_726477\|2020 | ···················· | ···················· | ····················· | ···················· | ···················· | ···················· |
| Y | 3181 | hCoV-19/Wales/PHWC-495D6F/2020\|EPI_ISL_726478\|2020 | ···················· | ······A············· | ····················· | ···················· | ···················· | ···················· |
| Y | 3182 | hCoV-19/Wales/PHWC-495D7E/2020\|EPI_ISL_726479\|2020 | ···················· | ···················· | ····················· | ···················· | ···················· | ···················· |
| Y | 3183 | hCoV-19/Wales/PHWC-495D8D/2020\|EPI_ISL_726480\|2020 | ···················· | ···················· | ····················· | ···················· | ···················· | ···················· |
| Y | 3184 | hCoV-19/Wales/PHWC-495D9C/2020\|EPI_ISL_726481\|2020 | ···················· | ···················· | ····················· | ···················· | ···················· | ···················· |
| Y | 3185 | hCoV-19/Wales/PHWC-495DAB/2020\|EPI_ISL_726482\|2020 | ···················· | ······A············· | ····················· | ···················· | ···················· | ···················· |
| Y | 3186 | hCoV-19/Wales/PHWC-495DC9/2020\|EPI_ISL_726484\|2020 | ···················· | ······A············· | ····················· | ···················· | ···················· | ···················· |
| Y | 3187 | hCoV-19/Wales/PHWC-495DD8/2020\|EPI_ISL_726485\|2020 | ···················· | ···················· | ····················· | ···················· | ···················· | ···················· |
| Y | 3188 | hCoV-19/Wales/PHWC-495DE7/2020\|EPI_ISL_726486\|2020 | ···················· | ···················· | ····················· | ···················· | ···················· | ···················· |
| Y | 3189 | hCoV-19/Wales/PHWC-495DF6/2020\|EPI_ISL_726487\|2020 | ···················· | ···················· | ····················· | ···················· | ···················· | ···················· |
| Y | 3190 | hCoV-19/Wales/PHWC-495E02/2020\|EPI_ISL_726488\|2020 | ···················· | ···················· | ····················· | ···················· | ···················· | ···················· |
| Y | 3191 | hCoV-19/Wales/PHWC-495E11/2020\|EPI_ISL_726489\|2020 | ···················· | ···················· | ····················· | ···················· | ···················· | ···················· |
| Y | 3192 | hCoV-19/England/PORT-2E98D6/2020\|EPI_ISL_727740\|20 | ···················· | ···················· | ····················· | ···················· | ······T············· | ···················· |
| Y | 3193 | hCoV-19/England/PORT-2E9997/2020\|EPI_ISL_727751\|20 | ···················· | ···················· | ····················· | ···················· | ···················· | ···················· |
| Y | 3194 | hCoV-19/England/PORT-2E9A1C/2020\|EPI_ISL_727755\|20 | ···················· | ·A·················· | ····················· | ···················· | ···················· | ···A················ |
| Y | 3195 | hCoV-19/England/PORT-2EAA2A/2020\|EPI_ISL_727764\|20 | ···················· | ···················· | ····················· | ···················· | ···················· | ···················· |
| Y | 3196 | hCoV-19/England/PORT-2EAA39/2020\|EPI_ISL_727765\|20 | ···················· | ···················· | ····················· | ···················· | ···················· | ···················· |
| Y | 3197 | hCoV-19/England/PORT-2EAA93/2020\|EPI_ISL_727767\|20 | ···················· | ···················· | ····················· | ···················· | ···················· | ···················· |
| Y | 3198 | hCoV-19/England/PORT-2EAAA2/2020\|EPI_ISL_727768\|20 | ···················· | ···················· | ····················· | ···················· | ···················· | ···················· |
| Y | 3199 | hCoV-19/England/PORT-2EAAEE/2020\|EPI_ISL_727771\|20 | ···················· | ···················· | ····················· | ···················· | ···················· | ···················· |
| Y | 3200 | hCoV-19/England/PORT-2EAAFD/2020\|EPI_ISL_727772\|20 | ···················· | ···················· | ····················· | ···················· | ···················· | ···················· |
| Y | 3201 | hCoV-19/England/PORT-2EAB27/2020\|EPI_ISL_727773\|20 | ···················· | ···················· | ····················· | ···················· | ···················· | ···················· |
| Y | 3202 | hCoV-19/England/QEUH-A9D271/2020\|EPI_ISL_727774\|20 | ···················· | ···················· | ····················· | ···················· | ···················· | ···················· |
| Y | 3203 | hCoV-19/England/QEUH-A9D29F/2020\|EPI_ISL_727776\|20 | ···················· | ···················· | ····················· | ···················· | ···················· | ···················· |
| Y | 3204 | hCoV-19/England/QEUH-A9D666/2020\|EPI_ISL_727781\|20 | ···················· | ···················· | ····················· | ···················· | ···················· | ···················· |
| Y | 3205 | hCoV-19/England/QEUH-A9D693/2020\|EPI_ISL_727782\|20 | ···················· | ···················· | ····················· | ···················· | ···················· | ···················· |
| Y | 3206 | hCoV-19/England/QEUH-A9D6DF/2020\|EPI_ISL_727784\|20 | ···················· | ···················· | ····················· | ···················· | ···················· | ···················· |
| Y | 3207 | hCoV-19/England/QEUH-A9D781/2020\|EPI_ISL_727787\|20 | ···················· | ···················· | ····················· | ···················· | ···················· | ···················· |
| Y | 3208 | hCoV-19/England/QEUH-A9D88E/2020\|EPI_ISL_727791\|20 | ···················· | ···················· | ····················· | ···················· | ···················· | ···················· |
| Y | 3209 | hCoV-19/England/QEUH-A9D8AC/2020\|EPI_ISL_727792\|20 | ···················· | ···················· | ····················· | ···················· | ···················· | ···················· |
| Y | 3210 | hCoV-19/Scotland/QEUH-A9E137/2020\|EPI_ISL_727798\|2 | ···················· | ···················· | ····················· | ···················· | ···················· | ···················· |
| Y | 3211 | hCoV-19/Portugal/PT2034/2020\|EPI_ISL_732250\|2020-1 | ···················· | ···················· | ····················· | ···················· | ···················· | ···················· |
| Y | 3212 | hCoV-19/Portugal/PT2035/2020\|EPI_ISL_732251\|2020-1 | ···················· | ···················· | ····················· | ···················· | ···················· | ···················· |
| Y | 3213 | hCoV-19/Portugal/PT2036/2020\|EPI_ISL_732252\|2020-1 | ···················· | ···················· | ····················· | ···················· | ···················· | ···················· |
| Y | 3214 | hCoV-19/Portugal/PT2037/2020\|EPI_ISL_732253\|2020-1 | ···················· | ·········A·········· | ····················· | ···················· | ······T············· | ···················· |
| Y | 3215 | hCoV-19/Portugal/PT2039/2020\|EPI_ISL_732254\|2020-1 | ···················· | ···················· | ····················· | ···················· | ···················· | ···················· |
| Y | 3216 | hCoV-19/Portugal/PT2040/2020\|EPI_ISL_732255\|2020-1 | ···················· | ···················· | ····················· | ···················· | ···················· | ···················· |
| Y | 3217 | hCoV-19/Portugal/PT2041/2020\|EPI_ISL_732256\|2020-1 | ···················· | ···················· | ····················· | ···················· | ···················· | ···················· |
| Y | 3218 | hCoV-19/Portugal/PT2042/2020\|EPI_ISL_732257\|2020-1 | ···················· | ···················· | ····················· | ···················· | ···················· | ···················· |
| Y | 3219 | hCoV-19/Portugal/PT2043/2020\|EPI_ISL_732258\|2020-1 | ···················· | ···················· | ····················· | ···················· | ···················· | ···················· |
| Y | 3221 | hCoV-19/Portugal/PT2045/2020\|EPI_ISL_732260\|2020-1 | ···················· | ···················· | ····················· | ···················· | ···················· | ···················· |
| Y | 3222 | hCoV-19/Portugal/PT2046/2020\|EPI_ISL_732261\|2020-1 | ···················· | ···················· | ····················· | ···················· | ···················· | ···················· |
| Y | 3223 | hCoV-19/Portugal/PT2047/2020\|EPI_ISL_732262\|2020-1 | ···················· | ···················· | ····················· | ···················· | ···················· | ···················· |
| Y | 3224 | hCoV-19/Portugal/PT2048/2020\|EPI_ISL_732263\|2020-1 | ···················· | ···················· | ····················· | ···················· | ···················· | ···················· |
| Y | 3225 | hCoV-19/Portugal/PT2050/2020\|EPI_ISL_732264\|2020-1 | ···················· | ···················· | ····················· | ···················· | ···················· | ···················· |
| Y | 3226 | hCoV-19/Portugal/PT2051/2020\|EPI_ISL_732265\|2020-1 | ···················· | ·········A·········· | ····················· | ···················· | ······T············· | ···················· |
| Y | 3227 | hCoV-19/Portugal/PT2052/2020\|EPI_ISL_732266\|2020-1 | ···················· | ·········A·········· | ···········T········· | ···················· | ······T············· | ···················· |
| Y | 3228 | hCoV-19/Portugal/PT2053/2020\|EPI_ISL_732267\|2020-1 | ···················· | ···················· | ····················· | ···················· | ···················· | ···················· |
| Y | 3229 | hCoV-19/Portugal/PT2054/2020\|EPI_ISL_732268\|2020-1 | ···················· | ···················· | ····················· | ···················· | ···················· | ···················· |
| Y | 3230 | hCoV-19/Portugal/PT2055/2020\|EPI_ISL_732269\|2020-1 | ···················· | ···················· | ····················· | ···················· | ···················· | ···················· |
| Y | 3231 | hCoV-19/Portugal/PT2056/2020\|EPI_ISL_732270\|2020-1 | ···················· | ···················· | ····················· | ···················· | ···················· | ···················· |
| Y | 3232 | hCoV-19/Portugal/PT2057/2020\|EPI_ISL_732271\|2020-1 | ···················· | ···················· | ····················· | ···················· | ···················· | ···················· |
| Y | 3233 | hCoV-19/Portugal/PT2058/2020\|EPI_ISL_732272\|2020-1 | ···················· | ···················· | ····················· | ···················· | ···················· | ···················· |
| Y | 3234 | hCoV-19/Portugal/PT2059/2020\|EPI_ISL_732273\|2020-1 | ···················· | ···················· | ····················· | ···················· | ···················· | ···················· |
| Y | 3235 | hCoV-19/Portugal/PT2060/2020\|EPI_ISL_732274\|2020-1 | ···················· | ···················· | ····················· | ···················· | ···················· | ···················· |
| Y | 3236 | hCoV-19/Portugal/PT2061/2020\|EPI_ISL_732275\|2020-1 | ···················· | ···················· | ····················· | ···················· | ···················· | ···················· |
| Y | 3237 | hCoV-19/Portugal/PT2062/2020\|EPI_ISL_732276\|2020-1 | ···················· | ···················· | ····················· | ···················· | ···················· | ···················· |
| Y | 3238 | hCoV-19/Portugal/PT2065/2020\|EPI_ISL_732279\|2020-1 | ···················· | ···················· | ····················· | ···················· | ···················· | ···················· |
| Y | 3239 | hCoV-19/Portugal/PT2076/2020\|EPI_ISL_732290\|2020-1 | ···················· | ···················· | ····················· | ···················· | ···················· | ···················· |
| Y | 3240 | hCoV-19/Portugal/PT2077/2020\|EPI_ISL_732291\|2020-1 | ···················· | ···················· | ····················· | ···················· | ···················· | ···················· |
| Y | 3241 | hCoV-19/Portugal/PT2078/2020\|EPI_ISL_732292\|2020-1 | ···················· | ···················· | ····················· | ···················· | ···················· | ···················· |
| Y | 3242 | hCoV-19/Portugal/PT2079/2020\|EPI_ISL_732293\|2020-1 | ···················· | ···················· | ····················· | ···················· | ···················· | ···················· |
| Y | 3243 | hCoV-19/England/C200026732/2020\|EPI_ISL_733582\|202 | ···················· | ···················· | ····················· | ···················· | ···················· | ···················· |
| Y | 3244 | hCoV-19/England/C200026740/2020\|EPI_ISL_733583\|202 | ···················· | ···················· | ····················· | ···················· | ···················· | ···················· |
| Y | 3245 | hCoV-19/England/C200027424/2020\|EPI_ISL_733585\|202 | ···················· | ···················· | ····················· | ···················· | ···················· | ···················· |
| Y | 3246 | hCoV-19/England/C200028755/2020\|EPI_ISL_733586\|202 | ···················· | ···················· | ····················· | ···················· | ···················· | ···················· |
| Y | 3247 | hCoV-19/England/C200028758/2020\|EPI_ISL_733587\|202 | ···················· | ···················· | ····················· | ···················· | ···················· | ···················· |
| Y | 3248 | hCoV-19/England/C200030587/2020\|EPI_ISL_733588\|202 | ···················· | ···················· | ····················· | ···················· | ···················· | ···················· |
| Y | 3249 | hCoV-19/England/205082082/2020\|EPI_ISL_733589\|2020 | ···················· | ·A·················· | ····················· | ···················· | ···················· | ···················· |
| Y | 3250 | hCoV-19/England/205082085/2020\|EPI_ISL_733592\|2020 | ···················· | ···················· | ····················· | ···················· | ···················· | ···················· |
| Y | 3251 | hCoV-19/England/205082086/2020\|EPI_ISL_733593\|2020 | ···················· | ···················· | ····················· | ···················· | ···················· | ···················· |
| Y | 3252 | hCoV-19/England/CAMC-C2CE62/2020\|EPI_ISL_733596\|20 | ···················· | ···················· | ····················· | ···················· | ···················· | ···················· |
| Y | 3253 | hCoV-19/England/ALDP-C23214/2020\|EPI_ISL_733598\|20 | ···················· | ···················· | ····················· | ···················· | ···················· | ···················· |
| Y | 3254 | hCoV-19/Belgium/rega-0803757/2020\|EPI_ISL_734900\|2 | ···················· | ···················· | ····················· | ···················· | ···················· | ···················· |

| | | | | | | | |
|---|---|---|---|---|---|---|---|
| Y | 3255 hCoV-19/USA/MI-MDHHS-SC22597/2020\|EPI_ISL_737170\|2 | ···················· | ·············A······ | ····················· | ···················· | ···················· | ···················· |
| Y | 3256 hCoV-19/USA/MI-MDHHS-SC22598/2020\|EPI_ISL_737171\|2 | ···················· | ···················· | ····················· | ···················· | ···················· | ···················· |
| Y | 3257 hCoV-19/USA/MI-MDHHS-SC22599/2020\|EPI_ISL_737172\|2 | ···················· | ···················· | ····················· | ···················· | ···················· | ···················· |
| Y | 3258 hCoV-19/USA/MI-MDHHS-SC22605/2020\|EPI_ISL_737178\|2 | ···················· | ···················· | ····················· | ···················· | ···················· | ···················· |
| Y | 3259 hCoV-19/Belgium/rega-12141347/2020\|EPI_ISL_738400\| | ···················· | ···················· | ····················· | ···················· | ···················· | ···················· |
| Y | 3260 hCoV-19/Belgium/rega-12111348/2020\|EPI_ISL_738401\| | ···················· | ···················· | ····················· | ···················· | ···················· | ···················· |
| Y | 3261 hCoV-19/Belgium/rega-12111349/2020\|EPI_ISL_738402\| | ···················· | ···················· | ····················· | ···················· | ···················· | ···················· |
| Y | 3262 hCoV-19/Belgium/rega-12111350/2020\|EPI_ISL_738403\| | ···················· | ···················· | ····················· | ···················· | ···················· | ···················· |
| Y | 3263 hCoV-19/Belgium/rega-12111351/2020\|EPI_ISL_738404\| | ···················· | ···················· | ····················· | ···················· | ···················· | ···················· |
| Y | 3264 hCoV-19/Belgium/rega-12111352/2020\|EPI_ISL_738405\| | ···················· | ···················· | ····················· | ···················· | ···················· | ···················· |
| Y | 3265 hCoV-19/Belgium/rega-12141353/2020\|EPI_ISL_738406\| | ···················· | ···················· | ····················· | ···················· | ···················· | ···················· |
| Y | 3266 hCoV-19/Belgium/rega-12111354/2020\|EPI_ISL_738407\| | ···················· | ···················· | ····················· | ···················· | ···················· | ···················· |
| Y | 3267 hCoV-19/Belgium/rega-12111355/2020\|EPI_ISL_738408\| | ···················· | ···················· | ····················· | ···················· | ···················· | ···················· |
| Y | 3268 hCoV-19/Belgium/rega-12141356/2020\|EPI_ISL_738409\| | ···················· | ···················· | ····················· | ···················· | ···················· | ···················· |
| Y | 3269 hCoV-19/Belgium/rega-12111357/2020\|EPI_ISL_738410\| | ···················· | ···················· | ····················· | ···················· | ···················· | ···················· |
| Y | 3270 hCoV-19/Belgium/rega-12111358/2020\|EPI_ISL_738411\| | ···················· | ···················· | ····················· | ···················· | ···················· | ···················· |
| Y | 3271 hCoV-19/Belgium/rega-12141359/2020\|EPI_ISL_738412\| | ···················· | ···················· | ····················· | ···················· | ···················· | ···················· |
| Y | 3272 hCoV-19/Belgium/rega-10131360/2020\|EPI_ISL_738413\| | ···················· | ···················· | ····················· | ···················· | ···················· | ···················· |
| Y | 3273 hCoV-19/Belgium/rega-12081361/2020\|EPI_ISL_738414\| | ···················· | ···················· | ····················· | ···················· | ···················· | ···················· |
| Y | 3274 hCoV-19/Belgium/rega-12141362/2020\|EPI_ISL_738415\| | ···················· | ···················· | ····················· | ···················· | ···················· | ···················· |
| Y | 3275 hCoV-19/Belgium/rega-12111363/2020\|EPI_ISL_738416\| | ···················· | ···················· | ····················· | ···················· | ···················· | ···················· |
| Y | 3276 hCoV-19/Belgium/rega-12141364/2020\|EPI_ISL_738417\| | ···················· | NN·················· | ····················· | ···················· | ···················· | ···················· |
| Y | 3277 hCoV-19/Belgium/rega-12111365/2020\|EPI_ISL_738418\| | ···················· | ···················· | ····················· | ···················· | ···················· | ···················· |
| Y | 3278 hCoV-19/Belgium/rega-10141366/2020\|EPI_ISL_738419\| | ···················· | ···················· | ····················· | ···················· | ···················· | ···················· |
| Y | 3279 hCoV-19/Belgium/rega-12141367/2020\|EPI_ISL_738420\| | ···················· | ···················· | ····················· | ···················· | ···················· | ···················· |
| Y | 3280 hCoV-19/Belgium/rega-12111368/2020\|EPI_ISL_738421\| | ···················· | ···················· | ····················· | ···················· | ···················· | ···················· |
| Y | 3281 hCoV-19/Belgium/rega-12111369/2020\|EPI_ISL_738422\| | ···················· | ···················· | ····················· | ···················· | ···················· | ···················· |
| Y | 3282 hCoV-19/Belgium/rega-12111370/2020\|EPI_ISL_738423\| | ···················· | ···················· | ····················· | ···················· | ···················· | ···················· |
| Y | 3283 hCoV-19/Belgium/rega-12141371/2020\|EPI_ISL_738424\| | ···················· | ···················· | ····················· | ···················· | ···················· | ···················· |
| Y | 3284 hCoV-19/Belgium/rega-10141372/2020\|EPI_ISL_738425\| | ···················· | ···················· | ····················· | ···················· | ···················· | ···················· |
| Y | 3285 hCoV-19/Belgium/rega-12111373/2020\|EPI_ISL_738426\| | ···················· | ···················· | ····················· | ···················· | ···················· | ···················· |
| Y | 3286 hCoV-19/Belgium/rega-12141374/2020\|EPI_ISL_738427\| | ···················· | ···················· | ····················· | ···················· | ···················· | ···················· |
| Y | 3287 hCoV-19/Belgium/rega-12111375/2020\|EPI_ISL_738428\| | ···················· | ···················· | ····················· | ···················· | ···················· | ···················· |
| Y | 3288 hCoV-19/Belgium/rega-12111376/2020\|EPI_ISL_738429\| | ···················· | ···················· | ····················· | ···················· | ···················· | ···················· |
| Y | 3289 hCoV-19/Belgium/rega-12141377/2020\|EPI_ISL_738430\| | ···················· | ···················· | ····················· | ···················· | ···················· | ···················· |
| Y | 3290 hCoV-19/Belgium/rega-12091379/2020\|EPI_ISL_738431\| | ···················· | ···················· | ····················· | ···················· | ···················· | ···················· |
| Y | 3291 hCoV-19/Belgium/rega-12141380/2020\|EPI_ISL_738432\| | ···················· | ···················· | ····················· | ···················· | ···················· | ···················· |
| Y | 3292 hCoV-19/Belgium/rega-12111381/2020\|EPI_ISL_738433\| | ···················· | ···················· | ····················· | ···················· | ···················· | ···················· |
| Y | 3293 hCoV-19/Belgium/rega-12111382/2020\|EPI_ISL_738434\| | ···················· | ···················· | ····················· | ···················· | ···················· | ···················· |
| Y | 3294 hCoV-19/Belgium/rega-12141383/2020\|EPI_ISL_738435\| | ···················· | ···················· | ····················· | ···················· | ···················· | ···················· |
| Y | 3295 hCoV-19/Belgium/rega-10141384/2020\|EPI_ISL_738436\| | ···················· | ···················· | ····················· | ···················· | ···················· | ···················· |
| Y | 3296 hCoV-19/Belgium/rega-12091385/2020\|EPI_ISL_738437\| | ···················· | ···················· | ····················· | ···················· | ···················· | ···················· |
| Y | 3297 hCoV-19/Belgium/rega-12111386/2020\|EPI_ISL_738438\| | ···················· | ···················· | ····················· | ···················· | ···················· | ···················· |
| Y | 3298 hCoV-19/Belgium/rega-12141387/2020\|EPI_ISL_738439\| | ···················· | ···················· | ····················· | ···················· | ···················· | ···················· |
| Y | 3299 hCoV-19/Belgium/rega-12111388/2020\|EPI_ISL_738440\| | ···················· | ···················· | ····················· | ···················· | ···················· | ···················· |
| Y | 3300 hCoV-19/Belgium/rega-12141389/2020\|EPI_ISL_738441\| | ···················· | ···················· | ····················· | ···················· | ···················· | ···················· |
| Y | 3301 hCoV-19/Belgium/rega-10141390/2020\|EPI_ISL_738442\| | ···················· | ···················· | ····················· | ···················· | ···················· | ···················· |
| Y | 3302 hCoV-19/Belgium/rega-12111391/2020\|EPI_ISL_738443\| | ···················· | ···················· | ····················· | ···················· | ···················· | ···················· |
| Y | 3303 hCoV-19/Belgium/rega-12111392/2020\|EPI_ISL_738444\| | ···················· | ···················· | ····················· | ···················· | ···················· | ···················· |
| Y | 3304 hCoV-19/Belgium/rega-12111393/2020\|EPI_ISL_738445\| | ···················· | ···················· | ····················· | ···················· | ···················· | ···················· |
| Y | 3305 hCoV-19/Belgium/rega-12111394/2020\|EPI_ISL_738446\| | ···················· | ···················· | ····················· | ···················· | ···················· | ···················· |
| Y | 3306 hCoV-19/Belgium/rega-10141395/2020\|EPI_ISL_738447\| | ···················· | ···················· | ····················· | ···················· | ···················· | ···················· |
| Y | 3307 hCoV-19/Belgium/rega-10141396/2020\|EPI_ISL_738448\| | ···················· | ···················· | ····················· | ···················· | ···················· | ···················· |
| Y | 3308 hCoV-19/Belgium/rega-12111397/2020\|EPI_ISL_738449\| | ···················· | ···················· | ····················· | ···················· | ···················· | ···················· |
| Y | 3309 hCoV-19/Belgium/rega-12141398/2020\|EPI_ISL_738450\| | ···················· | ···················· | ····················· | ···················· | ···················· | ···················· |
| Y | 3310 hCoV-19/Belgium/rega-12111399/2020\|EPI_ISL_738451\| | ···················· | ···················· | ····················· | ···················· | ···················· | ···················· |
| Y | 3311 hCoV-19/Belgium/rega-12141400/2020\|EPI_ISL_738452\| | ···················· | ···················· | ····················· | ···················· | ···················· | ···················· |
| Y | 3312 hCoV-19/Belgium/rega-10141401/2020\|EPI_ISL_738453\| | ···················· | ···················· | ····················· | ···················· | ···················· | ···················· |
| Y | 3313 hCoV-19/Belgium/rega-12141402/2020\|EPI_ISL_738454\| | ···················· | ···················· | ····················· | ···················· | ···················· | ···················· |
| Y | 3314 hCoV-19/Belgium/rega-12141403/2020\|EPI_ISL_738455\| | ···················· | ···················· | ····················· | ···················· | ···················· | ···················· |
| Y | 3315 hCoV-19/Belgium/rega-12111404/2020\|EPI_ISL_738456\| | ···················· | ···················· | ····················· | ···················· | ···················· | ···················· |
| Y | 3316 hCoV-19/Belgium/rega-12111405/2020\|EPI_ISL_738457\| | ···················· | ···················· | ····················· | ···················· | ···················· | ···················· |
| Y | 3317 hCoV-19/Belgium/rega-12141406/2020\|EPI_ISL_738458\| | ···················· | ···················· | ····················· | ···················· | ···················· | ···················· |
| Y | 3318 hCoV-19/Belgium/rega-12141407/2020\|EPI_ISL_738459\| | ···················· | ···················· | ····················· | ···················· | ···················· | ···················· |
| Y | 3319 hCoV-19/Belgium/rega-12141408/2020\|EPI_ISL_738460\| | ···················· | ···················· | ····················· | ···················· | ···················· | ···················· |
| Y | 3320 hCoV-19/Belgium/rega-12141409/2020\|EPI_ISL_738461\| | ···················· | ···················· | ····················· | ···················· | ···················· | ···················· |
| Y | 3321 hCoV-19/Belgium/rega-12111410/2020\|EPI_ISL_738462\| | ···················· | ···················· | ····················· | ···················· | ···················· | ···················· |
| Y | 3322 hCoV-19/Belgium/rega-12111411/2020\|EPI_ISL_738463\| | ···················· | ···················· | ····················· | ···················· | ···················· | ···················· |
| Y | 3323 hCoV-19/Belgium/rega-12141412/2020\|EPI_ISL_738464\| | ···················· | ···················· | ····················· | ···················· | ···················· | ···················· |
| Y | 3324 hCoV-19/Belgium/rega-12121413/2020\|EPI_ISL_738465\| | ···················· | ···················· | ····················· | ···················· | ···················· | ···················· |
| Y | 3325 hCoV-19/Belgium/rega-12141414/2020\|EPI_ISL_738466\| | ···················· | ···················· | ····················· | ···················· | ···················· | ···················· |
| Y | 3326 hCoV-19/Belgium/rega-12141415/2020\|EPI_ISL_738467\| | ···················· | ···················· | ····················· | ···················· | ···················· | ···················· |
| Y | 3327 hCoV-19/Belgium/rega-12111416/2020\|EPI_ISL_738468\| | ···················· | ···················· | ····················· | ···················· | ···················· | ···················· |
| Y | 3328 hCoV-19/Belgium/rega-12061417/2020\|EPI_ISL_738469\| | ···················· | ···················· | ····················· | ···················· | ···················· | ···················· |
| Y | 3329 hCoV-19/England/NORW-F3E51/2020\|EPI_ISL_725160\|202 | ···················· | ···················· | ····················· | ···················· | ···················· | ···················· |
| Y | 3330 hCoV-19/England/NORW-F3E60/2020\|EPI_ISL_725161\|202 | ···················· | ···················· | ····················· | ···················· | ···················· | ···················· |
| Y | 3331 hCoV-19/England/NORW-F3E8E/2020\|EPI_ISL_725162\|202 | ···················· | ···················· | ····················· | ···················· | ···················· | ···················· |
| Y | 3332 hCoV-19/England/NORW-F3E9D/2020\|EPI_ISL_725163\|202 | ···················· | ···················· | ····················· | ···················· | ···················· | ···················· |
| Y | 3333 hCoV-19/England/NORW-F3ECA/2020\|EPI_ISL_725166\|202 | ···················· | ···················· | ····················· | ···················· | ···················· | ···················· |
| Y | 3334 hCoV-19/England/NORW-F3EE8/2020\|EPI_ISL_725167\|202 | ···················· | ···················· | ····················· | ···················· | ···················· | ···················· |

| | | | | | | | |
|---|---|---|---|---|---|---|---|
| Y | 3335 hCoV-19/England/NORW-F3F7C/2020\|EPI_ISL_725172\|202 | ···················· | ···················· | ···················· | ···················· | ···················· | ···················· |
| Y | 3336 hCoV-19/England/NORW-F3F9A/2020\|EPI_ISL_725174\|202 | ···················· | ···················· | ···················· | ···················· | ···················· | ···················· |
| Y | 3337 hCoV-19/England/NORW-F3FA9/2020\|EPI_ISL_725175\|202 | ···················· | ···················· | ···················· | ···················· | ···················· | ···················· |
| Y | 3338 hCoV-19/England/NORW-F3FC7/2020\|EPI_ISL_725177\|202 | ···················· | ···················· | ···················· | ···················· | ···················· | ···················· |
| Y | 3339 hCoV-19/England/NORW-F3FD6/2020\|EPI_ISL_725178\|202 | ···················· | ···················· | ···················· | ···················· | ···················· | ···················· |
| Y | 3340 hCoV-19/England/NORW-F3FE5/2020\|EPI_ISL_725179\|202 | ···················· | ···················· | ···················· | ···················· | ···················· | ···················· |
| Y | 3341 hCoV-19/England/NORW-F40D2/2020\|EPI_ISL_725190\|202 | ···················· | ···················· | ···················· | ···················· | ···················· | ···················· |
| Y | 3342 hCoV-19/England/NORW-F40F0/2020\|EPI_ISL_725191\|202 | ···················· | ···················· | ···················· | ···················· | ···················· | ···················· |
| Y | 3343 hCoV-19/England/NORW-F410C/2020\|EPI_ISL_725192\|202 | ···················· | ···················· | ···················· | ···················· | ···················· | ···················· |
| Y | 3344 hCoV-19/England/NORW-F411B/2020\|EPI_ISL_725193\|202 | ···················· | ···················· | ···················· | ···················· | ···················· | ···················· |
| Y | 3345 hCoV-19/England/NORW-F412A/2020\|EPI_ISL_725194\|202 | ···················· | ···················· | ···················· | ···················· | ···················· | ···················· |
| Y | 3346 hCoV-19/England/NORW-F4148/2020\|EPI_ISL_725195\|202 | ···················· | ···················· | ···················· | ···················· | ···················· | ···················· |
| Y | 3347 hCoV-19/England/NORW-F4157/2020\|EPI_ISL_725196\|202 | ···················· | ···················· | ···················· | ···················· | ···················· | ···················· |
| Y | 3349 hCoV-19/Wales/PHWC-495E3F/2020\|EPI_ISL_726491\|2020 | ···················· | ···················· | ···················· | ···················· | ···················· | ···················· |
| Y | 3350 hCoV-19/Wales/PHWC-495E4E/2020\|EPI_ISL_726492\|2020 | ···················· | ···················· | ···················· | ···················· | ···················· | ···················· |
| Y | 3352 hCoV-19/Wales/PHWC-495E6C/2020\|EPI_ISL_726494\|2020 | ···················· | ···················· | ···················· | ···················· | ···················· | ···················· |
| Y | 3354 hCoV-19/Wales/PHWC-495E8A/2020\|EPI_ISL_726496\|2020 | ···················· | ···················· | ···················· | ···················· | ···················· | ···················· |
| Y | 3355 hCoV-19/Wales/PHWC-495E99/2020\|EPI_ISL_726497\|2020 | ···················· | ···················· | ···················· | ···················· | ···················· | ···················· |
| Y | 3356 hCoV-19/Wales/PHWC-495EA8/2020\|EPI_ISL_726498\|2020 | ···················· | ···················· | ···················· | ···················· | ···················· | ···················· |
| Y | 3358 hCoV-19/England/ALDP-C39F32/2020\|EPI_ISL_731000\|20 | ···················· | ···················· | ···················· | ···················· | ···················· | ···················· |
| Y | 3359 hCoV-19/England/ALDP-C3DB2B/2020\|EPI_ISL_731002\|20 | ···················· | ···················· | ···················· | ···················· | ···················· | ···················· |
| Y | 3360 hCoV-19/England/ALDP-C39F23/2020\|EPI_ISL_731006\|20 | ···················· | ···················· | ···················· | ···················· | ···················· | ···················· |
| Y | 3361 hCoV-19/England/ALDP-C3DB3A/2020\|EPI_ISL_731007\|20 | ···················· | ···················· | ···················· | ···················· | ···················· | ···················· |
| Y | 3362 hCoV-19/England/ALDP-C3C52E/2020\|EPI_ISL_731009\|20 | ···················· | ···················· | ···················· | ···················· | ···················· | ···················· |
| Y | 3363 hCoV-19/England/ALDP-C3BA11/2020\|EPI_ISL_731011\|20 | ···················· | ···················· | ···················· | ···················· | ···················· | ···················· |
| Y | 3364 hCoV-19/England/ALDP-C3BBA5/2020\|EPI_ISL_731013\|20 | ···················· | ···················· | ···················· | ···················· | ···················· | ···················· |
| Y | 3365 hCoV-19/England/ALDP-C3C60D/2020\|EPI_ISL_731018\|20 | ···················· | ·A·················· | ···················· | ···················· | ···················· | ···················· |
| Y | 3366 hCoV-19/England/ALDP-C39CC2/2020\|EPI_ISL_731019\|20 | ···················· | ···················· | ···················· | ···················· | ···················· | ···················· |
| Y | 3367 hCoV-19/England/ALDP-C3C324/2020\|EPI_ISL_731021\|20 | ···················· | ···················· | ···················· | ···················· | ···················· | ···················· |
| Y | 3368 hCoV-19/England/ALDP-C3B7FC/2020\|EPI_ISL_731023\|20 | ···················· | ···················· | ···················· | ···················· | ···················· | ···················· |
| Y | 3369 hCoV-19/England/ALDP-C39EF9/2020\|EPI_ISL_731027\|20 | ···················· | ···················· | ···················· | ···················· | ···················· | ···················· |
| Y | 3370 hCoV-19/England/ALDP-C3B756/2020\|EPI_ISL_731028\|20 | ···················· | ···················· | ···················· | ···················· | ···················· | ···················· |
| Y | 3371 hCoV-19/England/ALDP-C3CF5D/2020\|EPI_ISL_731029\|20 | ···················· | ···················· | ···················· | ···················· | ···················· | ···················· |
| Y | 3372 hCoV-19/England/ALDP-C3CC74/2020\|EPI_ISL_731030\|20 | ···················· | ···················· | ···················· | ···················· | ···················· | ···················· |
| Y | 3373 hCoV-19/England/ALDP-C3C99B/2020\|EPI_ISL_731031\|20 | ···················· | ···················· | ···················· | ···················· | ···················· | ···················· |
| Y | 3374 hCoV-19/England/ALDP-C3BA4E/2020\|EPI_ISL_731032\|20 | ···················· | ···················· | ···················· | ···················· | ···················· | ···················· |
| Y | 3375 hCoV-19/England/ALDP-C3A2A1/2020\|EPI_ISL_731033\|20 | ···················· | ···················· | ···················· | ···················· | ···················· | ···················· |
| Y | 3376 hCoV-19/England/ALDP-C3C7DD/2020\|EPI_ISL_731036\|20 | ···················· | ···················· | ···················· | ···················· | ···················· | ···················· |
| Y | 3377 hCoV-19/England/ALDP-C3C351/2020\|EPI_ISL_731037\|20 | ···················· | ···················· | ···················· | ···················· | ···················· | ···················· |
| Y | 3378 hCoV-19/England/ALDP-C3C931/2020\|EPI_ISL_731038\|20 | ···················· | ···················· | ···················· | ···················· | ···················· | ···················· |
| Y | 3379 hCoV-19/England/ALDP-C3CCCF/2020\|EPI_ISL_731039\|20 | ···················· | ···················· | ···················· | ···················· | ···················· | ···················· |
| Y | 3380 hCoV-19/England/CAMC-C2CA9B/2020\|EPI_ISL_733604\|20 | ···················· | ···················· | ···················· | ···················· | ·····T·············· | ···················· |
| Y | 3381 hCoV-19/England/MILK-C2FC1A/2020\|EPI_ISL_733607\|20 | ···················· | ···················· | ···················· | ···················· | ···················· | ···················· |
| Y | 3382 hCoV-19/England/CAMC-C2CFC9/2020\|EPI_ISL_733610\|20 | ···················· | ···················· | ···················· | ···················· | ···················· | ···················· |
| Y | 3383 hCoV-19/England/ALDP-C22E6C/2020\|EPI_ISL_733611\|20 | ···················· | ···················· | ···················· | ···················· | ···················· | ···················· |
| Y | 3384 hCoV-19/England/ALDP-C23056/2020\|EPI_ISL_733613\|20 | ···················· | ···················· | ···················· | ···················· | ···················· | ···················· |
| Y | 3385 hCoV-19/England/CAMC-C264A2/2020\|EPI_ISL_733614\|20 | ···················· | ···················· | ···················· | ···················· | ···················· | ···················· |
| Y | 3386 hCoV-19/England/MILK-C2FB1D/2020\|EPI_ISL_733617\|20 | ···················· | ···················· | ···················· | ···················· | ···················· | ···················· |
| Y | 3387 hCoV-19/England/ALDP-C22FF0/2020\|EPI_ISL_733618\|20 | ···················· | ···················· | ···················· | ···················· | ···················· | ···················· |
| Y | 3388 hCoV-19/England/ALDP-C231AE/2020\|EPI_ISL_733621\|20 | ···················· | ···················· | ···················· | ···················· | ···················· | ···················· |
| Y | 3389 hCoV-19/England/ALDP-C231F9/2020\|EPI_ISL_733623\|20 | ···················· | ···················· | ···················· | ···················· | ···················· | ···················· |
| Y | 3390 hCoV-19/England/CAMC-C2D414/2020\|EPI_ISL_733626\|20 | ···················· | ·A·················· | ···················· | ···················· | ···················· | ···A················ |
| Y | 3391 hCoV-19/England/MILK-C2F96E/2020\|EPI_ISL_733628\|20 | ···················· | ···················· | ···················· | ···················· | ···················· | ···················· |
| Y | 3392 hCoV-19/England/MILK-C2F9AA/2020\|EPI_ISL_733630\|20 | ···················· | ···················· | ···················· | ···················· | ···················· | ···················· |
| Y | 3393 hCoV-19/England/ALDP-C231BD/2020\|EPI_ISL_733635\|20 | ···················· | ···················· | ···················· | ···················· | ···················· | ···················· |
| Y | 3394 hCoV-19/England/ALDP-C2319F/2020\|EPI_ISL_733638\|20 | ···················· | ···················· | ···················· | ···················· | ···················· | ···················· |
| Y | 3395 hCoV-19/England/ALDP-C23C70/2020\|EPI_ISL_733641\|20 | ···················· | ···················· | ···················· | ···················· | ···················· | ···················· |
| Y | 3396 hCoV-19/England/ALDP-C244B3/2020\|EPI_ISL_733642\|20 | ···················· | ···················· | ···················· | ···················· | ···················· | ···················· |
| Y | 3397 hCoV-19/England/ALDP-C23C8F/2020\|EPI_ISL_733643\|20 | ···················· | ···················· | ···················· | ···················· | ···················· | ···················· |
| Y | 3398 hCoV-19/England/MILK-C305B1/2020\|EPI_ISL_733644\|20 | ···················· | ···················· | ···················· | ···················· | ···················· | ···················· |
| Y | 3399 hCoV-19/England/ALDP-C23144/2020\|EPI_ISL_733647\|20 | ···················· | ···················· | ···················· | ···················· | ···················· | ···················· |
| Y | 3400 hCoV-19/England/MILK-C2FAF2/2020\|EPI_ISL_733648\|20 | ···················· | ···················· | ···················· | ···················· | ···················· | ···················· |
| Y | 3401 hCoV-19/England/MILK-C2F940/2020\|EPI_ISL_733652\|20 | ···················· | ···················· | ···················· | ···················· | ···················· | ···················· |
| Y | 3402 hCoV-19/England/CAMC-C2D371/2020\|EPI_ISL_733656\|20 | ···················· | ···················· | ···················· | ···················· | ···················· | ···················· |
| Y | 3403 hCoV-19/England/MILK-C2FA89/2020\|EPI_ISL_733657\|20 | ···················· | ···················· | ···················· | ···················· | ·····T·············· | ···················· |
| Y | 3404 hCoV-19/England/ALDP-C2301A/2020\|EPI_ISL_733659\|20 | ···················· | ···················· | ···················· | ···················· | ···················· | ···················· |
| Y | 3405 hCoV-19/England/MILK-C305A2/2020\|EPI_ISL_733664\|20 | ···················· | ···················· | ···················· | ···················· | ···················· | ···················· |
| Y | 3406 hCoV-19/England/MILK-C30566/2020\|EPI_ISL_733669\|20 | ···················· | ···················· | ···················· | ···················· | ·····T·············· | ···················· |
| Y | 3407 hCoV-19/England/CAMC-C2C8FB/2020\|EPI_ISL_733681\|20 | ···················· | ·A·················· | ···················· | ···················· | ···················· | ···················· |
| Y | 3408 hCoV-19/England/ALDP-C23250/2020\|EPI_ISL_733682\|20 | ···················· | ···················· | ···················· | ···················· | ···················· | ···················· |
| Y | 3409 hCoV-19/England/ALDP-C231CC/2020\|EPI_ISL_733683\|20 | ···················· | ···················· | ···················· | ···················· | ···················· | ···················· |
| Y | 3410 hCoV-19/England/ALDP-C231EA/2020\|EPI_ISL_733685\|20 | ···················· | ···················· | ···················· | ···················· | ···················· | ···················· |
| Y | 3411 hCoV-19/England/MILK-C2F99B/2020\|EPI_ISL_733687\|20 | ···················· | ···················· | ···················· | ···················· | ···················· | ···················· |
| Y | 3412 hCoV-19/England/ALDP-C22EC6/2020\|EPI_ISL_733693\|20 | ···················· | ···················· | ···················· | ···················· | ·····T·············· | ···················· |
| Y | 3413 hCoV-19/England/CAMC-C2CAE6/2020\|EPI_ISL_733694\|20 | ···················· | ·A·················· | ···················· | ···················· | ···················· | ···················· |
| Y | 3414 hCoV-19/England/ALDP-C23C61/2020\|EPI_ISL_733697\|20 | ···················· | ···················· | ···················· | ···················· | ···················· | ···················· |
| Y | 3415 hCoV-19/England/CAMC-C2C916/2020\|EPI_ISL_733698\|20 | ···················· | ···················· | ···················· | ···················· | ···················· | ···················· |
| Y | 3416 hCoV-19/England/ALDP-C2328D/2020\|EPI_ISL_733699\|20 | ···················· | ···················· | ···················· | ···················· | ···················· | ···················· |
| Y | 3417 hCoV-19/Belgium/rega-0803758/2020\|EPI_ISL_734901\|2 | ···················· | ···················· | ···················· | ···················· | ···················· | ···················· |
| Y | 3418 hCoV-19/Belgium/rega-0803759/2020\|EPI_ISL_734902\|2 | ···················· | ···················· | ···················· | ···················· | ···················· | ···················· |

Y 3419 hCoV-19/Belgium/rega-0803760/2020|EPI_ISL_734903|2 ···················· ···················· ···················· ···················· ···················· ····················
Y 3420 hCoV-19/Belgium/rega-0803765/2020|EPI_ISL_734904|2 ···················· ···················· ···················· ···················· ···················· ····················
Y 3421 hCoV-19/Belgium/rega-0803766/2020|EPI_ISL_734905|2 ···················· ·A·················· ···················· ···················· ···················· ····················
Y 3422 hCoV-19/Belgium/rega-0804719/2020|EPI_ISL_734906|2 ···················· ···················· ···················· ···················· ···················· ····················
Y 3423 hCoV-19/Belgium/rega-0804726/2020|EPI_ISL_734907|2 ···················· ···················· ···················· ···················· ···················· ····················
Y 3424 hCoV-19/Belgium/rega-0804728/2020|EPI_ISL_734908|2 ···················· ···················· ···················· ···················· ···················· ····················
Y 3425 hCoV-19/Belgium/rega-0804729/2020|EPI_ISL_734909|2 ···················· ···················· ···················· ···················· ···················· ····················
Y 3426 hCoV-19/Belgium/rega-0804731/2020|EPI_ISL_734910|2 ···················· ···················· ···················· ···················· ···················· ····················
Y 3427 hCoV-19/Belgium/rega-0804735/2020|EPI_ISL_734911|2 ···················· ···················· ···················· ···················· ···················· ····················
Y 3428 hCoV-19/Belgium/rega-0804738/2020|EPI_ISL_734912|2 ···················· ···················· ···················· ···················· ···················· ····················
Y 3429 hCoV-19/Belgium/rega-0804742/2020|EPI_ISL_734913|2 ···················· ···················· ···················· ···················· ···················· ····················
Y 3430 hCoV-19/Belgium/rega-0804743/2020|EPI_ISL_734914|2 ···················· ···················· ···················· ···················· ···················· ····················
Y 3431 hCoV-19/Belgium/rega-0804744/2020|EPI_ISL_734915|2 ···················· ···················· ···················· ···················· ···················· ····················
Y 3432 hCoV-19/Belgium/rega-0804748/2020|EPI_ISL_734916|2 ···················· ···················· ···················· ···················· ···················· ····················
Y 3433 hCoV-19/Belgium/rega-0908933/2020|EPI_ISL_734917|2 ···················· ···················· ···················· ···················· ···················· ····················
Y 3434 hCoV-19/Belgium/rega-0909929/2020|EPI_ISL_734918|2 ···················· ···················· ···················· ···················· ···················· ····················
Y 3435 hCoV-19/Belgium/rega-0909930/2020|EPI_ISL_734919|2 ···················· ···················· ···················· ···················· ···················· ····················
Y 3436 hCoV-19/Belgium/rega-0909931/2020|EPI_ISL_734920|2 ···················· ···················· ···················· ···················· ···················· ····················
Y 3437 hCoV-19/Belgium/rega-0909932/2020|EPI_ISL_734921|2 ···················· ···················· ···················· ···················· ···················· ····················
Y 3438 hCoV-19/Belgium/rega-0909934/2020|EPI_ISL_734922|2 ···················· ···················· ···················· ···················· ···················· ····················
Y 3439 hCoV-19/Belgium/rega-0910935/2020|EPI_ISL_734923|2 ···················· ···················· ···················· ···················· ···················· ····················
Y 3440 hCoV-19/Belgium/rega-0911936/2020|EPI_ISL_734924|2 ···················· ···················· ···················· ···················· ···················· ····················
Y 3441 hCoV-19/Belgium/rega-0911937/2020|EPI_ISL_734925|2 ···················· ···················· ···················· ···················· ···················· ····················
Y 3442 hCoV-19/Belgium/rega-0911939/2020|EPI_ISL_734926|2 ···················· ···················· ···················· ···················· ···················· ····················
Y 3443 hCoV-19/Belgium/rega-0911940/2020|EPI_ISL_734927|2 ···················· ···················· ···················· ···················· ···················· ····················
Y 3444 hCoV-19/Belgium/rega-0911945/2020|EPI_ISL_734928|2 ···················· ···················· ···················· ···················· ···················· ····················
Y 3445 hCoV-19/Belgium/rega-0912938/2020|EPI_ISL_734929|2 ···················· ···················· ···················· ···················· ···················· ····················
Y 3446 hCoV-19/Belgium/rega-0912943/2020|EPI_ISL_734930|2 ···················· ···················· ···················· ···················· ···················· ····················
Y 3447 hCoV-19/Belgium/rega-0912944/2020|EPI_ISL_734931|2 ···················· ···················· ···················· ···················· ···················· ····················
Y 3448 hCoV-19/Belgium/rega-0913941/2020|EPI_ISL_734932|2 ···················· ···················· ···················· ···················· ···················· ····················
Y 3449 hCoV-19/Belgium/rega-0913942/2020|EPI_ISL_734933|2 ···················· ···················· ···················· ···················· ···················· ····················
Y 3450 hCoV-19/Belgium/rega-0914947/2020|EPI_ISL_734934|2 ···················· ···················· ···················· ···················· ···················· ····················
Y 3451 hCoV-19/Belgium/rega-09151217/2020|EPI_ISL_734935| ···················· ···················· ···················· ···················· ···················· ····················
Y 3452 hCoV-19/Belgium/rega-09151223/2020|EPI_ISL_734936| ···················· ···················· ···················· ···················· ···················· ····················
Y 3453 hCoV-19/Belgium/rega-0915946/2020|EPI_ISL_734937|2 ···················· ···················· ···················· ···················· ···················· ····················
Y 3454 hCoV-19/Belgium/rega-0915948/2020|EPI_ISL_734938|2 ···················· ···················· ···················· ···················· ···················· ····················
Y 3455 hCoV-19/Belgium/rega-0915949/2020|EPI_ISL_734939|2 ···················· ···················· ···················· ···················· ···················· ····················
Y 3456 hCoV-19/Belgium/rega-0915950/2020|EPI_ISL_734940|2 ···················· ···················· ···················· ···················· ···················· ····················
Y 3457 hCoV-19/Belgium/rega-0915951/2020|EPI_ISL_734941|2 ···················· ···················· ···················· ···················· ···················· ····················
Y 3458 hCoV-19/Belgium/rega-0915952/2020|EPI_ISL_734942|2 ···················· ···················· ···················· ···················· ···················· ····················
Y 3459 hCoV-19/Belgium/rega-0915953/2020|EPI_ISL_734943|2 ···················· ···················· ···················· ···················· ···················· ····················
Y 3460 hCoV-19/Belgium/rega-0915954/2020|EPI_ISL_734944|2 ···················· ···················· ···················· ···················· ···················· ····················
Y 3461 hCoV-19/Belgium/rega-0915955/2020|EPI_ISL_734945|2 ···················· ···················· ···················· ···················· ···················· ····················
Y 3462 hCoV-19/Belgium/rega-0915956/2020|EPI_ISL_734946|2 ···················· ···················· ···················· ···················· ···················· ····················
Y 3463 hCoV-19/Belgium/rega-0915957/2020|EPI_ISL_734947|2 ···················· ···················· ···················· ···················· ···················· ····················
Y 3464 hCoV-19/Belgium/rega-0915964/2020|EPI_ISL_734948|2 ···················· ···················· ···················· ···················· ···················· ····················
Y 3465 hCoV-19/Belgium/rega-0915965/2020|EPI_ISL_734949|2 ···················· ···················· ···················· ···················· ···················· ····················
Y 3466 hCoV-19/Belgium/rega-0915966/2020|EPI_ISL_734950|2 ···················· ···················· ···················· ···················· ···················· ····················
Y 3467 hCoV-19/Belgium/rega-0916958/2020|EPI_ISL_734951|2 ···················· ···················· ···················· ···················· ···················· ····················
Y 3468 hCoV-19/Belgium/rega-0916959/2020|EPI_ISL_734952|2 ···················· ···················· ···················· ···················· ···················· ····················
Y 3469 hCoV-19/Belgium/rega-0916960/2020|EPI_ISL_734953|2 ···················· ···················· ···················· ···················· ···················· ····················
Y 3470 hCoV-19/Belgium/rega-0916961/2020|EPI_ISL_734954|2 ···················· ···················· ···················· ···················· ···················· ····················
Y 3471 hCoV-19/Belgium/rega-0916962/2020|EPI_ISL_734955|2 ···················· ···················· ···················· ···················· ···················· ····················
Y 3472 hCoV-19/Belgium/rega-0916963/2020|EPI_ISL_734956|2 ···················· ···················· ···················· ···················· ···················· ····················
Y 3473 hCoV-19/Belgium/rega-0916967/2020|EPI_ISL_734957|2 ···················· ···················· ···················· ···················· ···················· ····················
Y 3474 hCoV-19/Belgium/rega-0916968/2020|EPI_ISL_734958|2 ···················· ···················· ···················· ···················· ···················· ····················
Y 3475 hCoV-19/Belgium/rega-0916969/2020|EPI_ISL_734959|2 ···················· ···················· ···················· ···················· ···················· ····················
Y 3476 hCoV-19/Belgium/rega-0916970/2020|EPI_ISL_734960|2 ···················· ···················· ···················· ···················· ···················· ····················
Y 3477 hCoV-19/Belgium/rega-0916971/2020|EPI_ISL_734961|2 ···················· ···················· ···················· ···················· ···················· ····················
Y 3478 hCoV-19/Belgium/rega-0916972/2020|EPI_ISL_734962|2 ···················· ···················· ···················· ···················· ···················· ····················
Y 3479 hCoV-19/Belgium/rega-0916973/2020|EPI_ISL_734963|2 ···················· ···················· ···················· ···················· ···················· ····················
Y 3480 hCoV-19/Belgium/rega-0916974/2020|EPI_ISL_734964|2 ···················· ···················· ···················· ···················· ···················· ····················
Y 3481 hCoV-19/Belgium/rega-0916975/2020|EPI_ISL_734965|2 ···················· ···················· ···················· ···················· ···················· ····················
Y 3482 hCoV-19/Belgium/rega-0916976/2020|EPI_ISL_734966|2 ···················· ···················· ···················· ···················· ···················· ····················
Y 3483 hCoV-19/Belgium/rega-0916977/2020|EPI_ISL_734967|2 ···················· ···················· ···················· ···················· ···················· ····················
Y 3484 hCoV-19/Belgium/rega-0916978/2020|EPI_ISL_734968|2 ···················· ···················· ···················· ···················· ···················· ····················
Y 3485 hCoV-19/Belgium/rega-0916979/2020|EPI_ISL_734969|2 ···················· ···················· ···················· ···················· ···················· ····················
Y 3486 hCoV-19/Belgium/rega-0916980/2020|EPI_ISL_734970|2 ···················· ···················· ···················· ···················· ···················· ····················
Y 3487 hCoV-19/Belgium/rega-0916981/2020|EPI_ISL_734971|2 ···················· ···················· ···················· ···················· ···················· ····················
Y 3488 hCoV-19/Belgium/rega-0916982/2020|EPI_ISL_734972|2 ···················· ···················· ···················· ···················· ···················· ····················
Y 3489 hCoV-19/Belgium/rega-09171048/2020|EPI_ISL_734973| ···················· ···················· ···················· ···················· ···················· ····················
Y 3490 hCoV-19/Belgium/rega-09171052/2020|EPI_ISL_734974| ···················· ···················· ···················· ···················· ···················· ····················
Y 3491 hCoV-19/Belgium/rega-0917983/2020|EPI_ISL_734975|2 ···················· ···················· ···················· ···················· ······T············· ····················
Y 3492 hCoV-19/Belgium/rega-0917984/2020|EPI_ISL_734976|2 ···················· ···················· ···················· ···················· ···················· ····················
Y 3493 hCoV-19/Belgium/rega-0917985/2020|EPI_ISL_734977|2 ···················· ···················· ···················· ···················· ···················· ····················
Y 3494 hCoV-19/Belgium/rega-0917986/2020|EPI_ISL_734978|2 ···················· ···················· ···················· ···················· ···················· ····················
Y 3495 hCoV-19/Belgium/rega-0917987/2020|EPI_ISL_734979|2 ···················· ···················· ···················· ···················· ···················· ····················
Y 3496 hCoV-19/Belgium/rega-0917988/2020|EPI_ISL_734980|2 ···················· ···················· ···················· ···················· ···················· ····················
Y 3497 hCoV-19/Belgium/rega-0917989/2020|EPI_ISL_734981|2 ···················· ···················· ···················· ···················· ···················· ····················
Y 3498 hCoV-19/Belgium/rega-0917990/2020|EPI_ISL_734982|2 ···················· ···················· ···················· ···················· ···················· ····················

Y 3499 hCoV-19/Belgium/rega-0917991/2020|EPI_ISL_734983|2 ···················· ·A·················· ···················· ···················· ···················· ····················
Y 3500 hCoV-19/Belgium/rega-0917992/2020|EPI_ISL_734984|2 ···················· ···················· ···················· ···················· ···················· ····················
Y 3501 hCoV-19/Belgium/rega-0917993/2020|EPI_ISL_734985|2 ···················· ···················· ···················· ···················· ···················· ····················
Y 3502 hCoV-19/Belgium/rega-0917994/2020|EPI_ISL_734986|2 ···················· ···················· ···················· ···················· ···················· ····················
Y 3503 hCoV-19/Belgium/rega-0917995/2020|EPI_ISL_734987|2 ···················· ···················· ···················· ···················· ···················· ····················
Y 3504 hCoV-19/Belgium/rega-0917997/2020|EPI_ISL_734988|2 ···················· ···················· ···················· ···················· ···················· ····················
Y 3505 hCoV-19/Belgium/rega-0917998/2020|EPI_ISL_734989|2 ···················· ···················· ···················· ···················· ···················· ····················
Y 3506 hCoV-19/Belgium/rega-09181049/2020|EPI_ISL_734992| ···················· ···················· ···················· ···················· ···················· ····················
Y 3507 hCoV-19/Belgium/rega-09181050/2020|EPI_ISL_734993| ···················· ···················· ···················· ···················· ···················· ····················
Y 3508 hCoV-19/Belgium/rega-09181051/2020|EPI_ISL_734994| ···················· ···················· ···················· ···················· ···················· ····················
Y 3509 hCoV-19/Belgium/rega-09181054/2020|EPI_ISL_734996| ···················· ···················· ···················· ···················· ···················· ····················
Y 3510 hCoV-19/Belgium/rega-09181055/2020|EPI_ISL_734997| ···················· ···················· ···················· ···················· ···················· ····················
Y 3511 hCoV-19/Belgium/rega-09181056/2020|EPI_ISL_734998| ···················· ···················· ···················· ···················· ···················· ····················
Y 3512 hCoV-19/Belgium/rega-09181057/2020|EPI_ISL_734999| ···················· ···················· ···················· ···················· ···················· ····················
Y 3513 hCoV-19/USA/MI-MDHHS-SC22579/2020|EPI_ISL_737153|2 ···················· ···················· ···················· ···················· ···················· ····················
Y 3514 hCoV-19/USA/MI-MDHHS-SC22580/2020|EPI_ISL_737154|2 ···················· ···················· ···················· ···················· ···················· ····················
Y 3515 hCoV-19/USA/MI-MDHHS-SC22581/2020|EPI_ISL_737155|2 ···················· ·············A······ ···················· ···················· ···················· ····················
Y 3516 hCoV-19/USA/MI-MDHHS-SC22582/2020|EPI_ISL_737156|2 ···················· ···················· ···················· ···················· ···················· ····················
Y 3517 hCoV-19/USA/MI-MDHHS-SC22583/2020|EPI_ISL_737157|2 ···················· ···················· ···················· ···················· ···················· ····················
Y 3518 hCoV-19/USA/MI-MDHHS-SC22585/2020|EPI_ISL_737158|2 ···················· ···················· ···················· ···················· ···················· ····················
Y 3519 hCoV-19/USA/MI-MDHHS-SC22586/2020|EPI_ISL_737159|2 ···················· ···················· ···················· ···················· ···················· ····················
Y 3520 hCoV-19/USA/MI-MDHHS-SC22587/2020|EPI_ISL_737160|2 ···················· ·············A······ ···················· ···················· ···················· ····················
Y 3521 hCoV-19/USA/MI-MDHHS-SC22588/2020|EPI_ISL_737161|2 ···················· ···················· ···················· ···················· ···················· ····················
Y 3522 hCoV-19/USA/MI-MDHHS-SC22589/2020|EPI_ISL_737162|2 ···················· ···················· ···················· ···················· ···················· ····················
Y 3523 hCoV-19/USA/MI-MDHHS-SC22590/2020|EPI_ISL_737163|2 ···················· ···················· ···················· ···················· ···················· ····················
Y 3524 hCoV-19/USA/MI-MDHHS-SC22591/2020|EPI_ISL_737164|2 ···················· ···················· ···················· ···················· ···················· ····················
Y 3525 hCoV-19/USA/MI-MDHHS-SC22592/2020|EPI_ISL_737165|2 ···················· ···················· ···················· ···················· ···················· ····················
Y 3526 hCoV-19/USA/MI-MDHHS-SC22593/2020|EPI_ISL_737166|2 ···················· ···················· ···················· ···················· ···················· ····················
Y 3527 hCoV-19/USA/MI-MDHHS-SC22594/2020|EPI_ISL_737167|2 ···················· ···················· ···················· ···················· ···················· ····················
Y 3528 hCoV-19/USA/MI-MDHHS-SC22595/2020|EPI_ISL_737168|2 ···················· ···················· ···················· ···················· ···················· ····················
Y 3529 hCoV-19/USA/MI-MDHHS-SC22596/2020|EPI_ISL_737169|2 ···················· ···················· ···················· ···················· ···················· ····················
Y 3530 hCoV-19/USA/MI-MDHHS-SC22600/2020|EPI_ISL_737173|2 ···················· ···················· ···················· ···················· ···················· ····················
Y 3531 hCoV-19/USA/MI-MDHHS-SC22601/2020|EPI_ISL_737174|2 ···················· ···················· ···················· ···················· ···················· ····················
Y 3533 hCoV-19/USA/MI-MDHHS-SC22603/2020|EPI_ISL_737176|2 ···················· ···················· ···················· ···················· ···················· ····················
Y 3534 hCoV-19/USA/MI-MDHHS-SC22604/2020|EPI_ISL_737177|2 ···················· ···················· ···················· ···················· ···················· ····················
Y 3535 hCoV-19/USA/MI-MDHHS-SC22606/2020|EPI_ISL_737179|2 ···················· ···················· ···················· ···················· ···················· ····················
Y 3536 hCoV-19/USA/MI-MDHHS-SC22607/2020|EPI_ISL_737180|2 ···················· ···················· ···················· ···················· ···················· ····················
Y 3537 hCoV-19/USA/MI-MDHHS-SC22608/2020|EPI_ISL_737181|2 ···················· ···················· ···················· ···················· ···················· ····················
Y 3538 hCoV-19/USA/MI-MDHHS-SC22609/2020|EPI_ISL_737182|2 ···················· ······A······A······ ···················· ···················· ···················· ····················
Y 3539 hCoV-19/USA/MI-MDHHS-SC22610/2020|EPI_ISL_737183|2 ···················· ···················· ···················· ···················· ···················· ····················
Y 3540 hCoV-19/USA/MI-MDHHS-SC22611/2020|EPI_ISL_737184|2 ···················· ···················· ···················· ···················· ···················· ····················
Y 3541 hCoV-19/USA/MI-MDHHS-SC22612/2020|EPI_ISL_737185|2 ···················· ···················· ···················· ···················· ···················· ····················
Y 3542 hCoV-19/USA/MI-MDHHS-SC22613/2020|EPI_ISL_737186|2 ···················· ···················· ···················· ···················· ···················· ····················
Y 3543 hCoV-19/USA/MI-MDHHS-SC22614/2020|EPI_ISL_737187|2 ···················· ···················· ···················· ···················· ···················· ····················
Y 3544 hCoV-19/USA/MI-MDHHS-SC22615/2020|EPI_ISL_737188|2 ········T··········· ·············A······ ···················· ···················· ···················· ····················
Y 3545 hCoV-19/USA/MI-MDHHS-SC22616/2020|EPI_ISL_737189|2 ···················· ···················· ···················· ···················· ···················· ····················
Y 3546 hCoV-19/USA/MI-MDHHS-SC22617/2020|EPI_ISL_737190|2 ···················· ······A······A······ ···················· ···················· ···················· ····················
Y 3547 hCoV-19/USA/MI-MDHHS-SC22618/2020|EPI_ISL_737191|2 ···················· ···················· ···················· ···················· ···················· ····················
Y 3548 hCoV-19/USA/MI-MDHHS-SC22619/2020|EPI_ISL_737192|2 ···················· ···················· ···················· ···················· ···················· ····················
Y 3549 hCoV-19/USA/MI-MDHHS-SC22620/2020|EPI_ISL_737193|2 ···················· ···················· ···················· ···················· ···················· ····················
Y 3550 hCoV-19/USA/MI-MDHHS-SC22621/2020|EPI_ISL_737194|2 ···················· ···················· ···················· ···················· ···················· ····················
Y 3551 hCoV-19/USA/MI-MDHHS-SC22622/2020|EPI_ISL_737195|2 ···················· ···················· ···················· ···················· ···················· ····················
Y 3552 hCoV-19/USA/MI-MDHHS-SC22623/2020|EPI_ISL_737196|2 ···················· ···················· ···················· ···················· ···················· ····················
Y 3553 hCoV-19/USA/MI-MDHHS-SC22646/2020|EPI_ISL_737197|2 ···················· ·············A······ ···················· ···················· ···················· ····················
Y 3554 hCoV-19/USA/MI-MDHHS-SC22647/2020|EPI_ISL_737198|2 ···················· ···················· ···················· ···················· ···················· ····················
Y 3555 hCoV-19/Belgium/rega-10131418/2020|EPI_ISL_738470| ···················· ···················· ···················· ···················· ···················· ····················
Y 3556 hCoV-19/Belgium/rega-12141419/2020|EPI_ISL_738471| ···················· ···················· ···················· ···················· ···················· ····················
Y 3557 hCoV-19/Belgium/rega-12141420/2020|EPI_ISL_738472| ···················· ···················· ···················· ···················· ···················· ····················
Y 3558 hCoV-19/Belgium/rega-12111421/2020|EPI_ISL_738473| ···················· ···················· ···················· ···················· ···················· ····················
Y 3559 hCoV-19/Belgium/rega-12141422/2020|EPI_ISL_738474| ···················· ···················· ···················· ···················· ···················· ····················
Y 3560 hCoV-19/Belgium/rega-10131423/2020|EPI_ISL_738475| ···················· ···················· ···················· ···················· ···················· ····················
Y 3561 hCoV-19/Belgium/rega-12141424/2020|EPI_ISL_738476| ···················· ···················· ···················· ···················· ···················· ····················
Y 3562 hCoV-19/Belgium/rega-12141425/2020|EPI_ISL_738477| ···················· ···················· ···················· ···················· ···················· ····················
Y 3563 hCoV-19/Belgium/rega-12111426/2020|EPI_ISL_738478| ···················· ···················· ···················· ···················· ···················· ····················
Y 3564 hCoV-19/Belgium/rega-12141427/2020|EPI_ISL_738479| ···················· ···················· ···················· ···················· ···················· ····················
Y 3565 hCoV-19/Belgium/rega-10141428/2020|EPI_ISL_738480| ···················· ···················· ···················· ···················· ···················· ····················
Y 3566 hCoV-19/Belgium/rega-12111429/2020|EPI_ISL_738481| ···················· ···················· ···················· ···················· ···················· ····················
Y 3567 hCoV-19/Belgium/rega-12141430/2020|EPI_ISL_738482| ···················· ···················· ···················· ···················· ···················· ····················
Y 3568 hCoV-19/Belgium/rega-12141431/2020|EPI_ISL_738483| ···················· ···················· ···················· ···················· ···················· ····················
Y 3569 hCoV-19/Belgium/rega-12111432/2020|EPI_ISL_738484| ···················· ···················· ···················· ···················· ···················· ····················
Y 3570 hCoV-19/Belgium/rega-12141433/2020|EPI_ISL_738485| ···················· ···················· ···················· ···················· ···················· ····················
Y 3571 hCoV-19/Belgium/rega-10131434/2020|EPI_ISL_738486| ···················· ···················· ···················· ···················· ···················· ····················
Y 3572 hCoV-19/Belgium/rega-12111435/2020|EPI_ISL_738487| ···················· ···················· ···················· ···················· ···················· ····················
Y 3573 hCoV-19/Belgium/rega-12141436/2020|EPI_ISL_738488| ···················· ···················· ···················· ···················· ···················· ····················
Y 3574 hCoV-19/Belgium/rega-12141437/2020|EPI_ISL_738489| ···················· ···················· ···················· ···················· ···················· ····················
Y 3575 hCoV-19/Belgium/rega-12111438/2020|EPI_ISL_738490| ···················· ···················· ···················· ···················· ···················· ····················
Y 3576 hCoV-19/Belgium/rega-12141439/2020|EPI_ISL_738491| ···················· ···················· ···················· ···················· ···················· ····················
Y 3577 hCoV-19/Belgium/rega-10131440/2020|EPI_ISL_738492| ···················· ···················· ···················· ···················· ···················· ····················
Y 3578 hCoV-19/Belgium/rega-12141441/2020|EPI_ISL_738493| ···················· ···················· ···················· ···················· ···················· ····················
Y 3579 hCoV-19/Belgium/rega-12141442/2020|EPI_ISL_738494| ···················· ···················· ···················· ···················· ···················· ····················

| | | | | | | | | |
|---|---|---|---|---|---|---|---|---|
| Y | 3580 | hCoV-19/Belgium/rega-12111443/2020\|EPI_ISL_738495\| | ···················· | ···················· | ····················· | ···················· | ···················· | ···················· |
| Y | 3581 | hCoV-19/Belgium/rega-12111444/2020\|EPI_ISL_738496\| | ···················· | ···················· | ····················· | ···················· | ···················· | ···················· |
| Y | 3582 | hCoV-19/Belgium/rega-12141445/2020\|EPI_ISL_738497\| | ···················· | ···················· | ····················· | ···················· | ···················· | ···················· |
| Y | 3583 | hCoV-19/Belgium/rega-10121446/2020\|EPI_ISL_738498\| | ···················· | ···················· | ····················· | ···················· | ···················· | ···················· |
| Y | 3584 | hCoV-19/Belgium/rega-12141447/2020\|EPI_ISL_738499\| | ···················· | ···················· | ····················· | ···················· | ···················· | ···················· |
| Y | 3585 | hCoV-19/England/NORW-F42AF/2020\|EPI_ISL_725205\|202 | ···················· | ···················· | ····················· | ···················· | ···················· | ···················· |
| Y | 3586 | hCoV-19/England/NORW-F437F/2020\|EPI_ISL_725211\|202 | ···················· | ···················· | ····················· | ···················· | ···················· | ···················· |
| Y | 3587 | hCoV-19/England/NORW-F43F7/2020\|EPI_ISL_725216\|202 | ···················· | ···················· | ····················· | ···················· | ···················· | ···················· |
| Y | 3588 | hCoV-19/England/NORW-F447C/2020\|EPI_ISL_725220\|202 | ···················· | ···················· | ····················· | ···················· | ···················· | ···················· |
| Y | 3589 | hCoV-19/England/NORW-F4746/2020\|EPI_ISL_725223\|202 | ···················· | ···················· | ····················· | ···················· | ···················· | ···················· |
| Y | 3590 | hCoV-19/England/NORW-F5156/2020\|EPI_ISL_725224\|202 | ···················· | ···················· | ····················· | ···················· | ···················· | ···················· |
| Y | 3591 | hCoV-19/England/NORW-F5183/2020\|EPI_ISL_725226\|202 | ···················· | ···················· | ····················· | ···················· | ···················· | ···················· |
| Y | 3592 | hCoV-19/England/NORW-F5192/2020\|EPI_ISL_725227\|202 | ···················· | ···················· | ····················· | ···················· | ···················· | ···················· |
| Y | 3593 | hCoV-19/England/NORW-F51A1/2020\|EPI_ISL_725228\|202 | ···················· | ···················· | ····················· | ···················· | ···················· | ···················· |
| Y | 3594 | hCoV-19/England/NORW-F51CF/2020\|EPI_ISL_725230\|202 | ···················· | ···················· | ····················· | ···················· | ···················· | ···················· |
| Y | 3595 | hCoV-19/England/NORW-F5208/2020\|EPI_ISL_725232\|202 | ···················· | ···················· | ····················· | ···················· | ···················· | ···················· |
| Y | 3596 | hCoV-19/England/NORW-F5226/2020\|EPI_ISL_725233\|202 | ···················· | ···················· | ····················· | ···················· | ···················· | ···················· |
| Y | 3597 | hCoV-19/England/NORW-F5262/2020\|EPI_ISL_725236\|202 | ···················· | ···················· | ····················· | ···················· | ···················· | ···················· |
| Y | 3598 | hCoV-19/England/NORW-F5271/2020\|EPI_ISL_725237\|202 | ···················· | ···················· | ····················· | ···················· | ···················· | ···················· |
| Y | 3599 | hCoV-19/England/NORW-F529F/2020\|EPI_ISL_725238\|202 | ···················· | ···················· | ····················· | ···················· | ···················· | ···················· |
| Y | 3600 | hCoV-19/England/NORW-F52AE/2020\|EPI_ISL_725239\|202 | ···················· | ···················· | ····················· | ···················· | ···················· | ···················· |
| Y | 3601 | hCoV-19/England/NORW-F52BD/2020\|EPI_ISL_725240\|202 | ···················· | ···················· | ····················· | ···················· | ···················· | ···················· |
| Y | 3602 | hCoV-19/England/NORW-F52CC/2020\|EPI_ISL_725241\|202 | ···················· | ···················· | ····················· | ···················· | ···················· | ···················· |
| Y | 3603 | hCoV-19/England/NORW-F5350/2020\|EPI_ISL_725246\|202 | ···················· | ···················· | ····················· | ···················· | ···················· | ···················· |
| Y | 3604 | hCoV-19/England/NORW-F538D/2020\|EPI_ISL_725248\|202 | ···················· | ···················· | ····················· | ···················· | ···················· | ···················· |
| Y | 3605 | hCoV-19/England/NORW-F539C/2020\|EPI_ISL_725249\|202 | ···················· | ···················· | ····················· | ···················· | ······T············· | ···················· |
| Y | 3606 | hCoV-19/England/NORW-F546C/2020\|EPI_ISL_725260\|202 | ···················· | ···················· | ····················· | ···················· | ···················· | ···················· |
| Y | 3607 | hCoV-19/England/NORW-F547B/2020\|EPI_ISL_725261\|202 | ···················· | ···················· | ····················· | ···················· | ···················· | ···················· |
| Y | 3608 | hCoV-19/England/NORW-F54A8/2020\|EPI_ISL_725264\|202 | ···················· | ···················· | ····················· | ···················· | ···················· | ···················· |
| Y | 3609 | hCoV-19/England/NORW-F54E4/2020\|EPI_ISL_725266\|202 | ···················· | ···················· | ····················· | ···················· | ···················· | ···················· |
| Y | 3610 | hCoV-19/England/NORW-F551E/2020\|EPI_ISL_725267\|202 | ···················· | ···················· | ····················· | ···················· | ···················· | ···················· |
| Y | 3611 | hCoV-19/England/NORW-F552D/2020\|EPI_ISL_725268\|202 | ···················· | ···················· | ····················· | ···················· | ···················· | ···················· |
| Y | 3612 | hCoV-19/England/NORW-F553C/2020\|EPI_ISL_725269\|202 | ···················· | ···················· | ····················· | ···················· | ···················· | ···················· |
| Y | 3613 | hCoV-19/England/NORW-F5569/2020\|EPI_ISL_725270\|202 | ···················· | ···················· | ····················· | ···················· | ···················· | ···················· |
| Y | 3614 | hCoV-19/England/NORW-F5596/2020\|EPI_ISL_725273\|202 | ···················· | ···················· | ····················· | ···················· | ···················· | ···················· |
| Y | 3615 | hCoV-19/England/NORW-F55A5/2020\|EPI_ISL_725274\|202 | ···················· | ···················· | ····················· | ···················· | ···················· | ···················· |
| Y | 3616 | hCoV-19/England/NORW-F55F0/2020\|EPI_ISL_725276\|202 | ···················· | ···················· | ····················· | ···················· | ···················· | ···················· |
| Y | 3617 | hCoV-19/England/NORW-F594F/2020\|EPI_ISL_725278\|202 | ···················· | ···················· | ····················· | ···················· | ···················· | ···················· |
| Y | 3618 | hCoV-19/England/NORW-F596D/2020\|EPI_ISL_725279\|202 | ···················· | ···················· | ····················· | ···················· | ···················· | ···················· |
| Y | 3619 | hCoV-19/Wales/PHWC-495EC6/2020\|EPI_ISL_726500\|2020 | ···················· | ···················· | ····················· | ···················· | ···················· | ···················· |
| Y | 3622 | hCoV-19/Wales/PHWC-495F0F/2020\|EPI_ISL_726504\|2020 | ···T················ | ···················· | ····················· | ···················· | ···················· | ···················· |
| Y | 3624 | hCoV-19/Wales/PHWC-495F3C/2020\|EPI_ISL_726507\|2020 | ···················· | ···················· | ····················· | ···················· | ···················· | ···················· |
| Y | 3626 | hCoV-19/Wales/PHWC-495F5A/2020\|EPI_ISL_726509\|2020 | ···················· | ···················· | ····················· | ···················· | ···················· | ···················· |
| Y | 3629 | hCoV-19/Wales/PHWC-495F96/2020\|EPI_ISL_726513\|2020 | ···················· | ···················· | ····················· | ···················· | ···················· | ···················· |
| Y | 3630 | hCoV-19/Wales/PHWC-495FC3/2020\|EPI_ISL_726516\|2020 | ···················· | ···················· | ····················· | ···················· | ···················· | ···················· |
| Y | 3632 | hCoV-19/Wales/PHWC-49600B/2020\|EPI_ISL_726518\|2020 | ···················· | ···················· | ····················· | ···················· | ···················· | ···················· |
| Y | 3633 | hCoV-19/Wales/PHWC-49601A/2020\|EPI_ISL_726519\|2020 | ···················· | ···················· | ····················· | ···················· | ···················· | ···················· |
| Y | 3634 | hCoV-19/Wales/PHWC-496029/2020\|EPI_ISL_726520\|2020 | ···················· | ···················· | ····················· | ···················· | ···················· | ···················· |
| Y | 3636 | hCoV-19/Wales/PHWC-496074/2020\|EPI_ISL_726524\|2020 | ···················· | ···················· | ····················· | ···················· | ···················· | ···················· |
| Y | 3639 | hCoV-19/Wales/PHWC-4960A1/2020\|EPI_ISL_726527\|2020 | ···················· | ···················· | ····················· | ···················· | ···················· | ···················· |
| Y | 3641 | hCoV-19/Wales/PHWC-4960CF/2020\|EPI_ISL_726529\|2020 | ···················· | ···················· | ····················· | ···················· | ···················· | ···················· |
| Y | 3643 | hCoV-19/Wales/PHWC-4960ED/2020\|EPI_ISL_726531\|2020 | ···················· | ···················· | ····················· | ···················· | ···················· | ···················· |
| Y | 3645 | hCoV-19/Wales/PHWC-496108/2020\|EPI_ISL_726533\|2020 | ···················· | ···················· | ····················· | ···················· | ···················· | ···················· |
| Y | 3646 | hCoV-19/Wales/PHWC-496117/2020\|EPI_ISL_726534\|2020 | ···················· | ···················· | ····················· | ···················· | ···················· | ···················· |
| Y | 3647 | hCoV-19/Wales/PHWC-496135/2020\|EPI_ISL_726536\|2020 | ···················· | ···················· | ····················· | ···················· | ···················· | ···················· |
| Y | 3648 | hCoV-19/Wales/PHWC-496144/2020\|EPI_ISL_726537\|2020 | ···················· | ···················· | ····················· | ···················· | ···················· | ···················· |
| Y | 3649 | hCoV-19/Wales/PHWC-496153/2020\|EPI_ISL_726538\|2020 | ···················· | ···················· | ····················· | ···················· | ···················· | ···················· |
| Y | 3650 | hCoV-19/Wales/PHWC-496162/2020\|EPI_ISL_726539\|2020 | ···················· | ···················· | ····················· | ···················· | ···················· | ···················· |
| Y | 3651 | hCoV-19/Wales/PHWC-496171/2020\|EPI_ISL_726540\|2020 | ···················· | ···················· | ····················· | ···················· | ···················· | ···················· |
| Y | 3652 | hCoV-19/Wales/PHWC-496180/2020\|EPI_ISL_726541\|2020 | ···················· | ···················· | ····················· | ···················· | ···················· | ···················· |
| Y | 3655 | hCoV-19/Wales/PHWC-4961DB/2020\|EPI_ISL_726544\|2020 | ···················· | ···················· | ····················· | ···················· | ···················· | ···················· |
| Y | 3656 | hCoV-19/Wales/PHWC-4961EA/2020\|EPI_ISL_726545\|2020 | ···················· | ···················· | ····················· | ···················· | ···················· | ···················· |
| Y | 3657 | hCoV-19/Wales/PHWC-4961F9/2020\|EPI_ISL_726546\|2020 | ···················· | ···················· | ····················· | ···················· | ···················· | ···················· |
| Y | 3658 | hCoV-19/Wales/PHWC-496205/2020\|EPI_ISL_726547\|2020 | ···················· | ···················· | ····················· | ···················· | ···················· | ···················· |
| Y | 3659 | hCoV-19/Wales/PHWC-496223/2020\|EPI_ISL_726549\|2020 | ···················· | ···················· | ····················· | ···················· | ···················· | ···················· |
| Y | 3660 | hCoV-19/Wales/PHWC-496232/2020\|EPI_ISL_726550\|2020 | ···················· | ···················· | ····················· | ···················· | ···················· | ···················· |
| Y | 3661 | hCoV-19/Wales/PHWC-496241/2020\|EPI_ISL_726551\|2020 | ···················· | ···················· | ····················· | ···················· | ···················· | ···················· |
| Y | 3663 | hCoV-19/Wales/PHWC-49627E/2020\|EPI_ISL_726553\|2020 | ···················· | ···················· | ····················· | ···················· | ···················· | ···················· |
| Y | 3664 | hCoV-19/Wales/PHWC-49628D/2020\|EPI_ISL_726554\|2020 | ···················· | ···················· | ····················· | ···················· | ···················· | ···················· |
| Y | 3665 | hCoV-19/Wales/PHWC-49629C/2020\|EPI_ISL_726555\|2020 | ···················· | ···················· | ····················· | ···················· | ···················· | ···················· |
| Y | 3667 | hCoV-19/Wales/PHWC-4962BA/2020\|EPI_ISL_726557\|2020 | ···················· | ···················· | ····················· | ···················· | ····T··············· | ···················· |
| Y | 3668 | hCoV-19/Wales/PHWC-4962C9/2020\|EPI_ISL_726558\|2020 | ···················· | ···················· | ····················· | ···················· | ···················· | ···················· |
| Y | 3669 | hCoV-19/Wales/PHWC-4962E7/2020\|EPI_ISL_726560\|2020 | ···················· | ···················· | ····················· | ···················· | ···················· | ···················· |
| Y | 3673 | hCoV-19/Wales/PHWC-49633F/2020\|EPI_ISL_726565\|2020 | ···················· | ···················· | ····················· | ···················· | ···················· | ···················· |
| Y | 3674 | hCoV-19/Wales/PHWC-49634E/2020\|EPI_ISL_726566\|2020 | ···················· | ···················· | ····················· | ···················· | ···················· | ···················· |
| Y | 3675 | hCoV-19/Wales/PHWC-49635D/2020\|EPI_ISL_726567\|2020 | ···················· | ···················· | ····················· | ···················· | ···················· | ···················· |
| Y | 3676 | hCoV-19/Wales/PHWC-49636C/2020\|EPI_ISL_726568\|2020 | ···················· | ···················· | ····················· | ···················· | ···················· | ···················· |
| Y | 3677 | hCoV-19/Wales/PHWC-49638A/2020\|EPI_ISL_726569\|2020 | ···················· | ···················· | ····················· | ···················· | ···················· | ···················· |
| Y | 3678 | hCoV-19/Wales/PHWC-496399/2020\|EPI_ISL_726570\|2020 | ···················· | ···················· | ····················· | ···················· | ···················· | ···················· |
| Y | 3679 | hCoV-19/Wales/PHWC-4963B7/2020\|EPI_ISL_726572\|2020 | ···················· | ···················· | ····················· | ···················· | ···················· | ···················· |

| | | | | | | | |
|---|---|---|---|---|---|---|---|
| Y | 3680 hCoV-19/Wales/PHWC-4963C6/2020\|EPI_ISL_726573\|2020 | ···················· | ···················· | ····················· | ···················· | ···················· | ···················· |
| Y | 3681 hCoV-19/Wales/PHWC-4963D5/2020\|EPI_ISL_726574\|2020 | ···················· | ···················· | ····················· | ···················· | ···················· | ···················· |
| Y | 3682 hCoV-19/Wales/PHWC-4963E4/2020\|EPI_ISL_726575\|2020 | ···················· | ···················· | ····················· | ···················· | ···················· | ···················· |
| Y | 3683 hCoV-19/Wales/PHWC-4963F3/2020\|EPI_ISL_726576\|2020 | ···················· | ···················· | ····················· | ···················· | ···················· | ···················· |
| Y | 3684 hCoV-19/Wales/PHWC-49640F/2020\|EPI_ISL_726577\|2020 | ···················· | ···················· | ····················· | ···················· | ···················· | ···················· |
| Y | 3685 hCoV-19/Wales/PHWC-49641E/2020\|EPI_ISL_726578\|2020 | ···················· | ···················· | ····················· | ···················· | ···················· | ···················· |
| Y | 3686 hCoV-19/Wales/PHWC-49642D/2020\|EPI_ISL_726579\|2020 | ···················· | ···················· | ····················· | ···················· | ···················· | ···················· |
| Y | 3689 hCoV-19/Wales/PHWC-4964F0/2020\|EPI_ISL_726592\|2020 | ···················· | ···················· | ····················· | ···················· | ···················· | ···················· |
| Y | 3690 hCoV-19/Wales/PHWC-49651B/2020\|EPI_ISL_726594\|2020 | ···················· | ···················· | ····················· | ···················· | ···················· | ···················· |
| Y | 3693 hCoV-19/Wales/PHWC-496548/2020\|EPI_ISL_726597\|2020 | ···················· | ···················· | ····················· | ···················· | ···················· | ···················· |
| Y | 3696 hCoV-19/England/QEUH-A9E182/2020\|EPI_ISL_727801\|20 | ···················· | ···················· | ····················· | ···················· | ···················· | ···················· |
| Y | 3697 hCoV-19/England/QEUH-A9E60B/2020\|EPI_ISL_727804\|20 | ···················· | ···················· | ····················· | ···················· | ···················· | ···················· |
| Y | 3698 hCoV-19/England/QEUH-A9E629/2020\|EPI_ISL_727805\|20 | ···················· | ···················· | ····················· | ···················· | ···················· | ···················· |
| Y | 3699 hCoV-19/England/QEUH-B7C63F/2020\|EPI_ISL_727807\|20 | ···················· | ···················· | ····················· | ···················· | ···················· | ···················· |
| Y | 3700 hCoV-19/England/QEUH-B7C65D/2020\|EPI_ISL_727808\|20 | ···················· | ···················· | ····················· | ···················· | ···················· | ···················· |
| Y | 3701 hCoV-19/England/QEUH-B7C67B/2020\|EPI_ISL_727809\|20 | ···················· | ···················· | ····················· | ···················· | ···················· | ···················· |
| Y | 3702 hCoV-19/England/QEUH-B7C68A/2020\|EPI_ISL_727810\|20 | ···················· | ···················· | ····················· | ···················· | ···················· | ···················· |
| Y | 3703 hCoV-19/England/QEUH-B7C699/2020\|EPI_ISL_727811\|20 | ···················· | ···················· | ····················· | ···················· | ···················· | ···················· |
| Y | 3704 hCoV-19/England/QEUH-B7C6B7/2020\|EPI_ISL_727812\|20 | ···················· | ···················· | ····················· | ···················· | ···················· | ···················· |
| Y | 3705 hCoV-19/England/QEUH-B7C6C6/2020\|EPI_ISL_727813\|20 | ···················· | ···················· | ····················· | ···················· | ···················· | ···················· |
| Y | 3706 hCoV-19/England/QEUH-B7C71E/2020\|EPI_ISL_727814\|20 | ···················· | ···················· | ····················· | ···················· | ···················· | ···················· |
| Y | 3707 hCoV-19/England/QEUH-B7C74B/2020\|EPI_ISL_727815\|20 | ···················· | ···················· | ····················· | ···················· | ···················· | ···················· |
| Y | 3708 hCoV-19/England/QEUH-B7C75A/2020\|EPI_ISL_727816\|20 | ···················· | ···················· | ····················· | ···················· | ···················· | ···················· |
| Y | 3709 hCoV-19/England/QEUH-B7C769/2020\|EPI_ISL_727817\|20 | ···················· | ···················· | ····················· | ···················· | ···················· | ···················· |
| Y | 3710 hCoV-19/England/QEUH-B7C787/2020\|EPI_ISL_727818\|20 | ···················· | ·A·················· | ····················· | ···················· | ···················· | ··A················· |
| Y | 3711 hCoV-19/England/QEUH-B7C796/2020\|EPI_ISL_727819\|20 | ···················· | ···················· | ····················· | ···················· | ···················· | ···················· |
| Y | 3712 hCoV-19/England/QEUH-B7C7B4/2020\|EPI_ISL_727820\|20 | ···················· | ···················· | ····················· | ···················· | ···················· | ··A················· |
| Y | 3713 hCoV-19/England/QEUH-B7C7C3/2020\|EPI_ISL_727821\|20 | ···················· | ···················· | ····················· | ···················· | ···················· | ···················· |
| Y | 3714 hCoV-19/England/QEUH-B7C7D2/2020\|EPI_ISL_727822\|20 | ···················· | ···················· | ····················· | ···················· | ···················· | ···················· |
| Y | 3715 hCoV-19/England/QEUH-B7C7E1/2020\|EPI_ISL_727823\|20 | ···················· | ···················· | ····················· | ···················· | ···················· | ···················· |
| Y | 3716 hCoV-19/England/QEUH-B7C7F0/2020\|EPI_ISL_727824\|20 | ···················· | ···················· | ····················· | ···················· | ···················· | ···················· |
| Y | 3717 hCoV-19/England/QEUH-B7C80C/2020\|EPI_ISL_727825\|20 | ···················· | ···················· | ····················· | ···················· | ···················· | ···················· |
| Y | 3718 hCoV-19/Scotland/QEUH-B7C81B/2020\|EPI_ISL_727826\|2 | ···················· | ···················· | ····················· | ···················· | ···················· | ···················· |
| Y | 3719 hCoV-19/England/QEUH-B7D56E/2020\|EPI_ISL_727827\|20 | ···················· | ···················· | ····················· | ···················· | ···················· | ···················· |
| Y | 3720 hCoV-19/England/QEUH-B7D8B0/2020\|EPI_ISL_727828\|20 | ···················· | ···················· | ····················· | ···················· | ···················· | ···················· |
| Y | 3721 hCoV-19/England/QEUH-B7D8CF/2020\|EPI_ISL_727829\|20 | ···················· | ···················· | ····················· | ···················· | ···················· | ···················· |
| Y | 3722 hCoV-19/England/QEUH-B7D8DE/2020\|EPI_ISL_727830\|20 | ···················· | ···················· | ····················· | ···················· | ···················· | ···················· |
| Y | 3723 hCoV-19/England/QEUH-B7D908/2020\|EPI_ISL_727831\|20 | ···················· | ···················· | ····················· | ···················· | ···················· | ···················· |
| Y | 3724 hCoV-19/England/QEUH-B7D917/2020\|EPI_ISL_727832\|20 | ···················· | ···················· | ····················· | ···················· | ···················· | ···················· |
| Y | 3725 hCoV-19/England/QEUH-B7D926/2020\|EPI_ISL_727833\|20 | ···················· | ···················· | ····················· | ···················· | ···················· | ···················· |
| Y | 3726 hCoV-19/England/QEUH-B7D944/2020\|EPI_ISL_727834\|20 | ···················· | ···················· | ····················· | ···················· | ···················· | ···················· |
| Y | 3727 hCoV-19/England/QEUH-B7D962/2020\|EPI_ISL_727835\|20 | ···················· | ···················· | ····················· | ···················· | ···················· | ···················· |
| Y | 3728 hCoV-19/England/QEUH-B7D971/2020\|EPI_ISL_727836\|20 | ···················· | ···················· | ····················· | ···················· | ···················· | ···················· |
| Y | 3729 hCoV-19/England/QEUH-B7D980/2020\|EPI_ISL_727837\|20 | ···················· | ···················· | ····················· | ···················· | ···················· | ···················· |
| Y | 3730 hCoV-19/England/QEUH-B7D9DB/2020\|EPI_ISL_727839\|20 | ···················· | ···················· | ····················· | ···················· | ···················· | ···················· |
| Y | 3731 hCoV-19/England/QEUH-B7D9EA/2020\|EPI_ISL_727840\|20 | ···················· | ···················· | ····················· | ···················· | ···················· | ···················· |
| Y | 3732 hCoV-19/England/QEUH-B7DA32/2020\|EPI_ISL_727841\|20 | ···················· | ···················· | ····················· | ···················· | ···················· | ···················· |
| Y | 3733 hCoV-19/England/QEUH-B7DA41/2020\|EPI_ISL_727842\|20 | ···················· | ···················· | ····················· | ···················· | ·····T·············· | ···················· |
| Y | 3734 hCoV-19/England/QEUH-B7DA50/2020\|EPI_ISL_727843\|20 | ···················· | ···················· | ····················· | ···················· | ···················· | ···················· |
| Y | 3735 hCoV-19/England/QEUH-B7DC2D/2020\|EPI_ISL_727844\|20 | ···················· | ·A·················· | ····················· | ···················· | ···················· | ···················· |
| Y | 3736 hCoV-19/England/QEUH-B7DCC3/2020\|EPI_ISL_727845\|20 | ···················· | ···················· | ····················· | ···················· | ···················· | ···················· |
| Y | 3737 hCoV-19/England/QEUH-B7DD0C/2020\|EPI_ISL_727846\|20 | ···················· | ···················· | ····················· | ···················· | ···················· | ···················· |
| Y | 3738 hCoV-19/England/QEUH-B7DD84/2020\|EPI_ISL_727847\|20 | ···················· | ···················· | ····················· | ···················· | ···················· | ···················· |
| Y | 3739 hCoV-19/England/QEUH-B7DDA2/2020\|EPI_ISL_727848\|20 | ···················· | ···················· | ····················· | ···················· | ···················· | ···················· |
| Y | 3740 hCoV-19/England/QEUH-B7E275/2020\|EPI_ISL_727849\|20 | ···················· | ···················· | ····················· | ···················· | ···················· | ···················· |
| Y | 3741 hCoV-19/England/QEUH-B7E2A2/2020\|EPI_ISL_727850\|20 | ···················· | ···················· | ····················· | ···················· | ···················· | ···················· |
| Y | 3742 hCoV-19/England/QEUH-B7E2C0/2020\|EPI_ISL_727851\|20 | ···················· | ···················· | ····················· | ···················· | ···················· | ···················· |
| Y | 3743 hCoV-19/England/QEUH-B7E2DF/2020\|EPI_ISL_727852\|20 | ···················· | ···················· | ····················· | ···················· | ···················· | ···················· |
| Y | 3744 hCoV-19/England/QEUH-B7E2EE/2020\|EPI_ISL_727853\|20 | ···················· | ···················· | ····················· | ···················· | ···················· | ···················· |
| Y | 3745 hCoV-19/England/QEUH-B7E309/2020\|EPI_ISL_727854\|20 | ···················· | ···················· | ····················· | ···················· | ···················· | ···················· |
| Y | 3746 hCoV-19/England/QEUH-B7E318/2020\|EPI_ISL_727855\|20 | ···················· | ···················· | ····················· | ···················· | ···················· | ···················· |
| Y | 3747 hCoV-19/England/QEUH-B7E327/2020\|EPI_ISL_727856\|20 | ···················· | ···················· | ····················· | ···················· | ···················· | ···················· |
| Y | 3748 hCoV-19/England/QEUH-B7E345/2020\|EPI_ISL_727858\|20 | ···················· | ···················· | ····················· | ···················· | ···················· | ···················· |
| Y | 3749 hCoV-19/England/QEUH-B7E363/2020\|EPI_ISL_727859\|20 | ···················· | ···················· | ····················· | ···················· | ···················· | ···················· |
| Y | 3750 hCoV-19/England/QEUH-B7E381/2020\|EPI_ISL_727860\|20 | ···················· | ···················· | ····················· | ···················· | ···················· | ···················· |
| Y | 3751 hCoV-19/England/SHEF-10C5DEF/2020\|EPI_ISL_727866\|2 | ···················· | ···················· | ····················· | ···················· | ···················· | ···················· |
| Y | 3752 hCoV-19/England/SHEF-10C614C/2020\|EPI_ISL_727867\|2 | ···················· | ···················· | ····················· | ···················· | ·····T·············· | ···················· |
| Y | 3753 hCoV-19/England/SHEF-10C62D0/2020\|EPI_ISL_727868\|2 | ···················· | ···················· | ····················· | ···················· | ···················· | ···················· |
| Y | 3754 hCoV-19/England/SHEF-10C771C/2020\|EPI_ISL_727870\|2 | ···················· | ···················· | ····················· | ···················· | ···················· | ···················· |
| Y | 3755 hCoV-19/England/SHEF-10C78BF/2020\|EPI_ISL_727872\|2 | ···················· | ···················· | ····················· | ···················· | ···················· | ···················· |
| Y | 3756 hCoV-19/England/SHEF-10C78EC/2020\|EPI_ISL_727873\|2 | ···················· | ···················· | ····················· | ···················· | ···················· | ···················· |
| Y | 3757 hCoV-19/England/SHEF-10C802F/2020\|EPI_ISL_727878\|2 | ···················· | ···················· | ····················· | ···················· | ···················· | ···················· |
| Y | 3758 hCoV-19/England/SHEF-10C8BB5/2020\|EPI_ISL_727883\|2 | ···················· | ···················· | ····················· | ···················· | ···················· | ···················· |
| Y | 3759 hCoV-19/England/SHEF-10C910D/2020\|EPI_ISL_727884\|2 | ···················· | ···················· | ····················· | ···················· | ···················· | ···················· |
| Y | 3760 hCoV-19/England/SHEF-10C92DD/2020\|EPI_ISL_727885\|2 | ···················· | ···················· | ····················· | ···················· | ···················· | ···················· |
| Y | 3761 hCoV-19/England/SHEF-10CAA89/2020\|EPI_ISL_727898\|2 | ···················· | ···················· | ····················· | ···················· | ···················· | ···················· |
| Y | 3762 hCoV-19/England/SHEF-10CAE41/2020\|EPI_ISL_727899\|2 | ···················· | ···················· | ····················· | ···················· | ···················· | ···················· |
| Y | 3763 hCoV-19/England/ALDP-C3D912/2020\|EPI_ISL_731004\|20 | ···················· | ···················· | ····················· | ···················· | ···················· | ···················· |
| Y | 3764 hCoV-19/England/ALDP-C39F05/2020\|EPI_ISL_731040\|20 | ···················· | ···················· | ····················· | ···················· | ···················· | ···················· |
| Y | 3765 hCoV-19/England/ALDP-C3C3D9/2020\|EPI_ISL_731041\|20 | ···················· | ···················· | ····················· | ···················· | ···················· | ···················· |

| | | | | | | | |
|---|---|---|---|---|---|---|---|
| Y | 3766 hCoV-19/England/ALDP-C3C47C/2020\|EPI_ISL_731042\|20 | ···················· | ···················· | ···················· | ···················· | ···················· | ··A················· |
| Y | 3767 hCoV-19/England/ALDP-C3D4A8/2020\|EPI_ISL_731044\|20 | ···················· | ···················· | ···················· | ···················· | ···················· | ···················· |
| Y | 3768 hCoV-19/England/ALDP-C3CF20/2020\|EPI_ISL_731046\|20 | ···················· | ···················· | ···················· | ···················· | ···················· | ···················· |
| Y | 3769 hCoV-19/England/ALDP-C3B932/2020\|EPI_ISL_731049\|20 | ···················· | ···················· | ···················· | ···················· | ···················· | ···················· |
| Y | 3770 hCoV-19/England/ALDP-C3D0A4/2020\|EPI_ISL_731051\|20 | ···················· | ···················· | ···················· | ···················· | ···················· | ···················· |
| Y | 3771 hCoV-19/England/ALDP-C39C1D/2020\|EPI_ISL_731052\|20 | ···················· | ···················· | ···················· | ···················· | ···················· | ···················· |
| Y | 3772 hCoV-19/England/ALDP-C3C825/2020\|EPI_ISL_731053\|20 | ···················· | ···················· | ···················· | ···················· | ···················· | ···················· |
| Y | 3773 hCoV-19/England/ALDP-C3A308/2020\|EPI_ISL_731054\|20 | ···················· | ···················· | ···················· | ···················· | ···················· | ···················· |
| Y | 3774 hCoV-19/England/ALDP-C3C342/2020\|EPI_ISL_731056\|20 | ···················· | ···················· | ···················· | ···················· | ···················· | ···················· |
| Y | 3775 hCoV-19/England/ALDP-C39F9C/2020\|EPI_ISL_731057\|20 | ···················· | ···················· | ···················· | ···················· | ···················· | ···················· |
| Y | 3776 hCoV-19/England/ALDP-C3C38E/2020\|EPI_ISL_731060\|20 | ···················· | ···················· | ···················· | ···················· | ···················· | ···················· |
| Y | 3777 hCoV-19/England/ALDP-C3C3AC/2020\|EPI_ISL_731061\|20 | ···················· | ···················· | ···················· | ···················· | ···················· | ···················· |
| Y | 3778 hCoV-19/England/ALDP-C39F14/2020\|EPI_ISL_731065\|20 | ···················· | ···················· | ···················· | ···················· | ···················· | ···················· |
| Y | 3779 hCoV-19/England/ALDP-C3A3EA/2020\|EPI_ISL_731066\|20 | ···················· | ···················· | ···················· | ···················· | ···················· | ···················· |
| Y | 3780 hCoV-19/England/ALDP-C3B97E/2020\|EPI_ISL_731067\|20 | ···················· | ···················· | ···················· | ···················· | ···················· | ···················· |
| Y | 3781 hCoV-19/England/ALDP-C3BD54/2020\|EPI_ISL_731069\|20 | ···················· | ···················· | ···················· | ···················· | ···················· | ···················· |
| Y | 3782 hCoV-19/England/ALDP-C3C7FB/2020\|EPI_ISL_731071\|20 | ···················· | ···················· | ···················· | ···················· | ···················· | ···················· |
| Y | 3783 hCoV-19/England/ALDP-C3CA3E/2020\|EPI_ISL_731072\|20 | ···················· | ···················· | ···················· | ···················· | ···················· | ···················· |
| Y | 3784 hCoV-19/England/ALDP-C3B7B0/2020\|EPI_ISL_731073\|20 | ···················· | ···················· | ···················· | ···················· | ···················· | ···················· |
| Y | 3785 hCoV-19/England/ALDP-C3A326/2020\|EPI_ISL_731074\|20 | ···················· | ···················· | ···················· | ···················· | ···················· | ···················· |
| Y | 3786 hCoV-19/England/ALDP-C3DB76/2020\|EPI_ISL_731075\|20 | ···················· | ···················· | ···················· | ···················· | ···················· | ···················· |
| Y | 3787 hCoV-19/England/ALDP-C3A344/2020\|EPI_ISL_731076\|20 | ···················· | ···················· | ···················· | ···················· | ···················· | ···················· |
| Y | 3788 hCoV-19/England/ALDP-C3CF99/2020\|EPI_ISL_731077\|20 | ···················· | ···················· | ···················· | ···················· | ···················· | ···················· |
| Y | 3789 hCoV-19/England/ALDP-C3C61C/2020\|EPI_ISL_731078\|20 | ···················· | ···················· | ···················· | ···················· | ···················· | ··A················· |
| Y | 3790 hCoV-19/England/ALDP-C3DB67/2020\|EPI_ISL_731081\|20 | ···················· | ···················· | ···················· | ···················· | ···················· | ···················· |
| Y | 3791 hCoV-19/England/ALDP-C39EAE/2020\|EPI_ISL_731082\|20 | ···················· | ···················· | ···················· | ···················· | ···················· | ···················· |
| Y | 3792 hCoV-19/England/ALDP-C3A441/2020\|EPI_ISL_731083\|20 | ···················· | ···················· | ···················· | ···················· | ···················· | ···················· |
| Y | 3793 hCoV-19/England/ALDP-C3CA2F/2020\|EPI_ISL_731085\|20 | ···················· | ···················· | ···················· | ···················· | ···················· | ···················· |
| Y | 3794 hCoV-19/England/ALDP-C3A256/2020\|EPI_ISL_731086\|20 | ···················· | ···················· | ···················· | ···················· | ···················· | ···················· |
| Y | 3795 hCoV-19/England/ALDP-C3C807/2020\|EPI_ISL_731089\|20 | ···················· | ···················· | ···················· | ···················· | ···················· | ···················· |
| Y | 3796 hCoV-19/England/ALDP-C3DB1C/2020\|EPI_ISL_731092\|20 | ···················· | ···················· | ···················· | ···················· | ···················· | ···················· |
| Y | 3797 hCoV-19/England/ALDP-C3B9BA/2020\|EPI_ISL_731093\|20 | ···················· | ···················· | ···················· | ···················· | ···················· | ···················· |
| Y | 3798 hCoV-19/England/ALDP-C3DE40/2020\|EPI_ISL_731095\|20 | ···················· | ···················· | ···················· | ···················· | ···················· | ···················· |
| Y | 3799 hCoV-19/England/ALDP-C3A49C/2020\|EPI_ISL_731096\|20 | ···················· | ···················· | ···················· | ···················· | ···················· | ···················· |
| Y | 3800 hCoV-19/England/ALDP-C3D4B7/2020\|EPI_ISL_731097\|20 | ···················· | ···················· | ···················· | ···················· | ···················· | ···················· |
| Y | 3801 hCoV-19/England/ALDP-C3C412/2020\|EPI_ISL_731098\|20 | ···················· | ···················· | ···················· | ···················· | ···················· | ···················· |
| Y | 3802 hCoV-19/England/MILK-C2FFE4/2020\|EPI_ISL_733666\|20 | ···················· | ···················· | ···················· | ···················· | ···················· | ···················· |
| Y | 3803 hCoV-19/Belgium/rega-0917999/2020\|EPI_ISL_734990\|2 | ···················· | ···················· | ···················· | ···················· | ···················· | ···················· |
| Y | 3804 hCoV-19/Belgium/rega-09181047/2020\|EPI_ISL_734991\| | ···················· | ·A·················· | ···················· | ···················· | ···················· | ···················· |
| Y | 3805 hCoV-19/Belgium/rega-09181053/2020\|EPI_ISL_734995\| | ···················· | ···················· | ···················· | ···················· | ···················· | ···················· |
| Y | 3806 hCoV-19/Nigeria/NCDC-4a/2020\|EPI_ISL_737201\|2020-0 | ···················· | ···················· | ···················· | ···················· | ···················· | ···················· |
| Y | 3807 hCoV-19/Finland/13ES21A9/2020\|EPI_ISL_737209\|2020- | ···················· | ·A·················· | ···················· | ···················· | ···················· | ··A················· |
| Y | 3809 hCoV-19/Finland/13KS15A3/2020\|EPI_ISL_737211\|2020- | ···················· | ···················· | ···················· | ···················· | ···················· | ···················· |
| Y | 3810 hCoV-19/Finland/13LS10AG11/2020\|EPI_ISL_737212\|202 | ···················· | ·A·················· | ···················· | ···················· | ···················· | ··A················· |
| Y | 3811 hCoV-19/Finland/13LS11AH1/2020\|EPI_ISL_737213\|2020 | ···················· | ·A·················· | ···················· | ···················· | ···················· | ··A················· |
| Y | 3812 hCoV-19/Finland/13LS12AG7/2020\|EPI_ISL_737214\|2020 | ···················· | ···················· | ···················· | ···················· | ···················· | ···················· |
| Y | 3813 hCoV-19/Finland/13LS12AH5/2020\|EPI_ISL_737215\|2020 | ···················· | ·A·················· | ···················· | ···················· | ···················· | ··A················· |
| Y | 3814 hCoV-19/Finland/13LS12AH8/2020\|EPI_ISL_737216\|2020 | ···················· | ···················· | ···················· | ···················· | ···················· | ···················· |
| Y | 3815 hCoV-19/Finland/13LS13AC1/2020\|EPI_ISL_737217\|2020 | ···················· | ·A·················· | ···················· | ···················· | ···················· | ··A················· |
| Y | 3816 hCoV-19/Finland/13LS13AF9/2020\|EPI_ISL_737218\|2020 | ···················· | ···················· | ···················· | ···················· | ···················· | ···················· |
| Y | 3817 hCoV-19/Finland/13LS14AC1/2020\|EPI_ISL_737219\|2020 | ···················· | ·A·················· | ···················· | ···················· | ···················· | ··A················· |
| Y | 3818 hCoV-19/Finland/13LS8AA5/2020\|EPI_ISL_737232\|2020- | ···················· | ···················· | ···················· | ···················· | ···················· | ···················· |
| Y | 3819 hCoV-19/Finland/13LS9AH11/2020\|EPI_ISL_737233\|2020 | ···················· | ···················· | ···················· | ···················· | ···················· | ···················· |
| Y | 3820 hCoV-19/USA/CA-LACPHL-AF00001/2020\|EPI_ISL_737236\| | ···················· | ···················· | ···················· | ···················· | ···················· | ···················· |
| Y | 3821 hCoV-19/USA/CA-LACPHL-AF00002/2020\|EPI_ISL_737237\| | ···················· | ···················· | ···················· | ···················· | ···················· | ···················· |
| Y | 3822 hCoV-19/USA/CA-LACPHL-AF00003/2020\|EPI_ISL_737238\| | ···················· | ···················· | ···················· | ···················· | ···················· | ···················· |
| Y | 3823 hCoV-19/USA/CA-LACPHL-AF00004/2020\|EPI_ISL_737239\| | ···················· | ·A····A············· | ···················· | ···················· | ···················· | ···················· |
| Y | 3824 hCoV-19/USA/CA-LACPHL-AF00005/2020\|EPI_ISL_737240\| | ···················· | ·A·················· | ···················· | ···················· | ···················· | ···················· |
| Y | 3825 hCoV-19/USA/CA-LACPHL-AF00006/2020\|EPI_ISL_737241\| | ···················· | ···················· | ···················· | ···················· | ···················· | ···················· |
| Y | 3826 hCoV-19/USA/CA-LACPHL-AF00008/2020\|EPI_ISL_737243\| | ···················· | ·A·················· | ···················· | ···················· | ···················· | ···················· |
| Y | 3827 hCoV-19/USA/CA-LACPHL-AF00009/2020\|EPI_ISL_737244\| | ···················· | ·A·················· | ···················· | ···················· | ···················· | ···················· |
| Y | 3828 hCoV-19/USA/CA-LACPHL-AF00010/2020\|EPI_ISL_737245\| | ···················· | ···················· | ···················· | ···················· | ···················· | ···················· |
| Y | 3829 hCoV-19/USA/CA-LACPHL-AF00011/2020\|EPI_ISL_737246\| | ···················· | ···················· | ···················· | ···················· | ···················· | ···················· |
| Y | 3830 hCoV-19/USA/CA-LACPHL-AF00012/2020\|EPI_ISL_737247\| | ···················· | ···················· | ···················· | ···················· | ···················· | ···················· |
| Y | 3831 hCoV-19/USA/CA-LACPHL-AF00013/2020\|EPI_ISL_737248\| | ···················· | ···················· | ···················· | ···················· | ···················· | ···················· |
| Y | 3832 hCoV-19/USA/CA-LACPHL-AF00014/2020\|EPI_ISL_737249\| | ···················· | ···················· | ···················· | ···················· | ···················· | ···················· |
| Y | 3833 hCoV-19/USA/CA-LACPHL-AF00016/2020\|EPI_ISL_737251\| | ···················· | ···················· | ···················· | ···················· | ···················· | ···················· |
| Y | 3834 hCoV-19/USA/CA-LACPHL-AF00017/2020\|EPI_ISL_737252\| | ···················· | ·············A······ | ···················· | ···················· | ···················· | ···················· |
| Y | 3835 hCoV-19/USA/CA-LACPHL-AF00018/2020\|EPI_ISL_737253\| | ···················· | ···················· | ···················· | ···················· | ···················· | ···················· |
| Y | 3836 hCoV-19/USA/CA-LACPHL-AF00019/2020\|EPI_ISL_737254\| | ···················· | ···················· | ···················· | ···················· | ···················· | ···················· |
| Y | 3837 hCoV-19/USA/CA-LACPHL-AF00020/2020\|EPI_ISL_737255\| | ···················· | ···················· | ···················· | ···················· | ···················· | ···················· |
| Y | 3838 hCoV-19/USA/CA-LACPHL-AF00021/2020\|EPI_ISL_737256\| | ···················· | ···················· | ···················· | ···················· | ···················· | ···················· |
| Y | 3839 hCoV-19/USA/CA-LACPHL-AF00023/2020\|EPI_ISL_737258\| | ···················· | ···················· | ···················· | ···················· | ···················· | ···················· |
| Y | 3840 hCoV-19/USA/CA-LACPHL-AF00024/2020\|EPI_ISL_737259\| | ···················· | ···················· | ···················· | ···················· | ···················· | ···················· |
| Y | 3841 hCoV-19/England/NORW-F53AB/2020\|EPI_ISL_725250\|202 | ···················· | ···················· | ···················· | ···················· | ···················· | ···················· |
| Y | 3842 hCoV-19/England/NORW-F53D8/2020\|EPI_ISL_725252\|202 | ···················· | ···················· | ···················· | ···················· | ···················· | ···················· |
| Y | 3843 hCoV-19/England/NORW-F53E7/2020\|EPI_ISL_725253\|202 | ···················· | ···················· | ···················· | ···················· | ···················· | ···················· |
| Y | 3844 hCoV-19/England/NORW-F5402/2020\|EPI_ISL_725255\|202 | ···················· | ···················· | ···················· | ···················· | ···················· | ···················· |
| Y | 3845 hCoV-19/England/NORW-F5411/2020\|EPI_ISL_725256\|202 | ···················· | ···················· | ···················· | ···················· | ···················· | ···················· |
| Y | 3846 hCoV-19/England/NORW-F598B/2020\|EPI_ISL_725281\|202 | ···················· | ···················· | ···················· | ···················· | ···················· | ···················· |

| | | | | | | | | |
|---|---|---|---|---|---|---|---|---|
| Y | 3847 | hCoV-19/England/NORW-F599A/2020\|EPI_ISL_725282\|202 | .................... | .................... | .................... | .................... | .................... | .................... |
| Y | 3848 | hCoV-19/England/NORW-F59E5/2020\|EPI_ISL_725285\|202 | .................... | .................... | .................... | .................... | .................... | .................... |
| Y | 3849 | hCoV-19/England/NORW-F5A2E/2020\|EPI_ISL_725288\|202 | .................... | .................... | .................... | .................... | .................... | .................... |
| Y | 3850 | hCoV-19/England/NORW-F5A5B/2020\|EPI_ISL_725289\|202 | .................... | .................... | .................... | .................... | .................... | .................... |
| Y | 3851 | hCoV-19/England/NORW-F5A79/2020\|EPI_ISL_725291\|202 | .................... | .................... | .................... | .................... | .................... | .................... |
| Y | 3852 | hCoV-19/England/NORW-F5A88/2020\|EPI_ISL_725292\|202 | .................... | .................... | .................... | .................... | .................... | .................... |
| Y | 3853 | hCoV-19/England/NORW-F5A97/2020\|EPI_ISL_725293\|202 | .................... | .................... | .................... | .................... | .................... | .................... |
| Y | 3854 | hCoV-19/England/NORW-F5AA6/2020\|EPI_ISL_725294\|202 | .................... | .................... | .................... | .................... | .................... | .................... |
| Y | 3855 | hCoV-19/England/NORW-F5AB5/2020\|EPI_ISL_725295\|202 | .................... | .................... | .................... | .................... | .................... | .................... |
| Y | 3856 | hCoV-19/England/NORW-F64C5/2020\|EPI_ISL_725296\|202 | .................... | .................... | .................... | .................... | .................... | .................... |
| Y | 3857 | hCoV-19/England/NORW-F64E3/2020\|EPI_ISL_725297\|202 | .................... | .................... | .................... | .................... | .................... | .................... |
| Y | 3858 | hCoV-19/England/NORW-F651D/2020\|EPI_ISL_725298\|202 | .................... | .................... | .................... | .................... | .................... | .................... |
| Y | 3859 | hCoV-19/England/NORW-F660B/2020\|EPI_ISL_725300\|202 | .................... | .................... | .................... | .................... | .................... | .................... |
| Y | 3860 | hCoV-19/England/NORW-F66ED/2020\|EPI_ISL_725301\|202 | .................... | .................... | .................... | .................... | .................... | .................... |
| Y | 3861 | hCoV-19/England/NORW-F6E21/2020\|EPI_ISL_725303\|202 | .................... | .................... | .................... | .................... | .................... | .................... |
| Y | 3863 | hCoV-19/England/NORW-F6E7C/2020\|EPI_ISL_725307\|202 | .................... | .................... | .................... | .................... | .................... | .................... |
| Y | 3864 | hCoV-19/England/NORW-F6E8B/2020\|EPI_ISL_725308\|202 | .................... | .................... | .................... | .................... | .................... | .................... |
| Y | 3865 | hCoV-19/England/NORW-F6E9A/2020\|EPI_ISL_725309\|202 | .................... | .................... | .................... | .................... | .................... | .................... |
| Y | 3866 | hCoV-19/England/NORW-F6EB8/2020\|EPI_ISL_725311\|202 | .................... | .................... | .................... | .................... | .................... | .................... |
| Y | 3867 | hCoV-19/England/NORW-F6F00/2020\|EPI_ISL_725315\|202 | .................... | .................... | .................... | .................... | .................... | .................... |
| Y | 3868 | hCoV-19/England/NORW-F6FE2/2020\|EPI_ISL_725318\|202 | .................... | .................... | .................... | .................... | .................... | .................... |
| Y | 3869 | hCoV-19/England/NORW-F702A/2020\|EPI_ISL_725319\|202 | .................... | .................... | .................... | .................... | .................... | .................... |
| Y | 3870 | hCoV-19/Wales/PHWC-49643C/2020\|EPI_ISL_726580\|2020 | .................... | .................... | .................... | .................... | .................... | .................... |
| Y | 3871 | hCoV-19/Wales/PHWC-49644B/2020\|EPI_ISL_726581\|2020 | .................... | .................... | .................... | .................... | .................... | .................... |
| Y | 3872 | hCoV-19/Wales/PHWC-49645A/2020\|EPI_ISL_726582\|2020 | .................... | .................... | .................... | .................... | .................... | .................... |
| Y | 3874 | hCoV-19/Wales/PHWC-496478/2020\|EPI_ISL_726584\|2020 | .................... | .................... | .................... | .................... | .................... | .................... |
| Y | 3875 | hCoV-19/Wales/PHWC-496487/2020\|EPI_ISL_726585\|2020 | .................... | .................... | .................... | .................... | .................... | .................... |
| Y | 3876 | hCoV-19/Wales/PHWC-496496/2020\|EPI_ISL_726586\|2020 | .................... | .................... | .................... | .................... | .................... | .................... |
| Y | 3877 | hCoV-19/Wales/PHWC-4964A5/2020\|EPI_ISL_726587\|2020 | .................... | .................... | .................... | .................... | .................... | .................... |
| Y | 3880 | hCoV-19/Wales/PHWC-496575/2020\|EPI_ISL_726600\|2020 | .................... | .................... | .................... | .................... | .................... | .................... |
| Y | 3882 | hCoV-19/Wales/PHWC-496593/2020\|EPI_ISL_726602\|2020 | .................... | .................... | .................... | .................... | .................... | .................... |
| Y | 3883 | hCoV-19/Wales/PHWC-4965A2/2020\|EPI_ISL_726603\|2020 | .................... | .................... | .................... | .................... | .................... | .................... |
| Y | 3886 | hCoV-19/Wales/PHWC-4965DF/2020\|EPI_ISL_726606\|2020 | .................... | .................... | .................... | .................... | .................... | .................... |
| Y | 3887 | hCoV-19/Wales/PHWC-496645/2020\|EPI_ISL_726613\|2020 | .................... | .................... | .................... | .................... | .................... | .................... |
| Y | 3888 | hCoV-19/Wales/PHWC-496654/2020\|EPI_ISL_726614\|2020 | .................... | .................... | .................... | .................... | .................... | .................... |
| Y | 3889 | hCoV-19/Wales/PHWC-496672/2020\|EPI_ISL_726616\|2020 | .................... | .................... | .................... | .................... | .................... | .................... |
| Y | 3890 | hCoV-19/Wales/PHWC-496681/2020\|EPI_ISL_726617\|2020 | .................... | .................... | .................... | .................... | .................... | .................... |
| Y | 3892 | hCoV-19/Wales/PHWC-496760/2020\|EPI_ISL_726631\|2020 | .................... | .................... | .................... | .................... | .................... | .................... |
| Y | 3893 | hCoV-19/Wales/PHWC-49679D/2020\|EPI_ISL_726634\|2020 | .................... | .................... | .................... | .................... | .................... | .................... |
| Y | 3894 | hCoV-19/Wales/PHWC-4967AC/2020\|EPI_ISL_726635\|2020 | .................... | .................... | .................... | .................... | .................... | .................... |
| Y | 3896 | hCoV-19/Wales/PHWC-496803/2020\|EPI_ISL_726640\|2020 | .................... | .................... | .................... | .................... | .................... | .................... |
| Y | 3898 | hCoV-19/Wales/PHWC-496821/2020\|EPI_ISL_726642\|2020 | .................... | .................... | .................... | .................... | .................... | .................... |
| Y | 3899 | hCoV-19/Wales/PHWC-496830/2020\|EPI_ISL_726643\|2020 | .................... | .................... | .................... | .................... | .................... | .................... |
| Y | 3902 | hCoV-19/Wales/PHWC-49687C/2020\|EPI_ISL_726647\|2020 | .................... | .................... | .................... | .................... | .................... | .................... |
| Y | 3904 | hCoV-19/Wales/PHWC-49689A/2020\|EPI_ISL_726649\|2020 | .................... | .................... | .................... | .................... | .................... | .................... |
| Y | 3905 | hCoV-19/England/SHEF-10CB5C3/2020\|EPI_ISL_727902\|2 | .................... | .................... | .................... | .................... | .................... | .................... |
| Y | 3906 | hCoV-19/England/SHEF-10CB639/2020\|EPI_ISL_727903\|2 | .................... | .................... | .................... | .................... | .................... | .................... |
| Y | 3907 | hCoV-19/England/SHEF-10CB8AC/2020\|EPI_ISL_727904\|2 | .................... | .................... | .................... | .................... | .................... | .................... |
| Y | 3908 | hCoV-19/England/SHEF-10CD761/2020\|EPI_ISL_727916\|2 | .................... | .................... | .................... | .................... | .................... | .................... |
| Y | 3909 | hCoV-19/England/SHEF-10CDEB7/2020\|EPI_ISL_727920\|2 | .................... | .................... | .................... | .................... | .................... | .................... |
| Y | 3910 | hCoV-19/England/SHEF-10CDEE4/2020\|EPI_ISL_727921\|2 | .................... | .................... | .................... | .................... | .................... | .................... |
| Y | 3911 | hCoV-19/England/SHEF-10CE645/2020\|EPI_ISL_727924\|2 | .................... | .................... | .................... | .................... | .................... | .................... |
| Y | 3912 | hCoV-19/England/SHEF-10CE9B5/2020\|EPI_ISL_727925\|2 | .................... | .................... | .................... | .................... | .................... | .................... |
| Y | 3913 | hCoV-19/England/SHEF-10CF125/2020\|EPI_ISL_727928\|2 | .................... | .................... | .................... | .................... | .................... | .................... |
| Y | 3914 | hCoV-19/England/SHEF-10CF222/2020\|EPI_ISL_727929\|2 | .................... | .................... | .................... | .................... | .................... | .................... |
| Y | 3915 | hCoV-19/England/SHEF-10CF389/2020\|EPI_ISL_727931\|2 | .................... | .................... | .................... | .................... | .................... | .................... |
| Y | 3916 | hCoV-19/England/SHEF-10CF7BA/2020\|EPI_ISL_727934\|2 | .................... | .................... | .................... | .................... | .................... | .................... |
| Y | 3917 | hCoV-19/England/SHEF-10D0F45/2020\|EPI_ISL_727942\|2 | .................... | .................... | .................... | .................... | .................... | .................... |
| Y | 3918 | hCoV-19/England/SHEF-10D10E7/2020\|EPI_ISL_727943\|2 | .................... | .................... | .................... | .................... | .................... | .................... |
| Y | 3919 | hCoV-19/England/SHEF-10D1542/2020\|EPI_ISL_727944\|2 | .................... | .................... | .................... | .................... | .................... | .................... |
| Y | 3920 | hCoV-19/England/SHEF-10D1991/2020\|EPI_ISL_727946\|2 | .................... | .................... | .................... | .................... | .................... | .................... |
| Y | 3921 | hCoV-19/England/SHEF-10D1A16/2020\|EPI_ISL_727947\|2 | .................... | .................... | .................... | .................... | .................... | .................... |
| Y | 3922 | hCoV-19/England/SHEF-10D3D75/2020\|EPI_ISL_727964\|2 | .................... | .................... | .................... | .................... | .................... | .................... |
| Y | 3923 | hCoV-19/England/SHEF-10D447F/2020\|EPI_ISL_727968\|2 | .................... | .................... | .................... | .................... | .................... | .................... |
| Y | 3924 | hCoV-19/England/SHEF-10D4503/2020\|EPI_ISL_727969\|2 | .................... | .................... | .................... | .................... | .................... | .................... |
| Y | 3925 | hCoV-19/England/SHEF-10D4A04/2020\|EPI_ISL_727970\|2 | .................... | .................... | .................... | .................... | .................... | .................... |
| Y | 3926 | hCoV-19/England/SHEF-10D53BD/2020\|EPI_ISL_727971\|2 | .................... | .................... | .................... | .................... | .................... | .................... |
| Y | 3927 | hCoV-19/England/SHEF-10D5696/2020\|EPI_ISL_727972\|2 | .................... | .................... | .................... | .................... | .................... | .................... |
| Y | 3928 | hCoV-19/England/SHEF-10D5DA0/2020\|EPI_ISL_727974\|2 | .................... | .................... | .................... | .................... | .................... | .................... |
| Y | 3929 | hCoV-19/England/SHEF-10D6097/2020\|EPI_ISL_727975\|2 | .................... | .................... | .................... | .................... | .................... | .................... |
| Y | 3930 | hCoV-19/England/ALDP-C3B7A1/2020\|EPI_ISL_731100\|20 | .................... | .................... | .................... | .................... | .................... | .................... |
| Y | 3931 | hCoV-19/England/ALDP-C3CF11/2020\|EPI_ISL_731102\|20 | .................... | .................... | .................... | .................... | .................... | .................... |
| Y | 3932 | hCoV-19/England/ALDP-C3D9A9/2020\|EPI_ISL_731103\|20 | .................... | .................... | .................... | .................... | .................... | .................... |
| Y | 3933 | hCoV-19/England/ALDP-C3A2CF/2020\|EPI_ISL_731104\|20 | .................... | .................... | .................... | .................... | .................... | .................... |
| Y | 3934 | hCoV-19/England/ALDP-C3A2B0/2020\|EPI_ISL_731106\|20 | .................... | .................... | .................... | .................... | .................... | .................... |
| Y | 3935 | hCoV-19/England/ALDP-C3C764/2020\|EPI_ISL_731108\|20 | .................... | .................... | .................... | .................... | .................... | .................... |
| Y | 3936 | hCoV-19/England/ALDP-C3CF3F/2020\|EPI_ISL_731109\|20 | .................... | .................... | .................... | .................... | .................... | .................... |
| Y | 3937 | hCoV-19/England/ALDP-C39E08/2020\|EPI_ISL_731110\|20 | .................... | .................... | .................... | .................... | .................... | .................... |
| Y | 3938 | hCoV-19/England/ALDP-C3B729/2020\|EPI_ISL_731112\|20 | .................... | .................... | .................... | .................... | .................... | .................... |
| Y | 3939 | hCoV-19/England/ALDP-C3CFB7/2020\|EPI_ISL_731113\|20 | .................... | .................... | .................... | .................... | .................... | .................... |

| | | | | | | | | |
|---|---|---|---|---|---|---|---|---|
| Y | 3940 hCoV-19/England/ALDP-C3B99C/2020\|EPI_ISL_731114\|20 | ···················· | ·A·················· | ···················· | ···················· | ···················· | ···A················ | |
| Y | 3941 hCoV-19/England/ALDP-C3CF02/2020\|EPI_ISL_731115\|20 | ···················· | ···················· | ···················· | ···················· | ···················· | ···A················ | |
| Y | 3942 hCoV-19/England/ALDP-C3C95F/2020\|EPI_ISL_731118\|20 | ···················· | ···················· | ···················· | ···················· | ···················· | ···················· | |
| Y | 3943 hCoV-19/England/ALDP-C39C0E/2020\|EPI_ISL_731120\|20 | ···················· | ···················· | ···················· | ···················· | ···················· | ···················· | |
| Y | 3944 hCoV-19/England/ALDP-C3D96D/2020\|EPI_ISL_731121\|20 | ···················· | ···················· | ···················· | ···················· | ···················· | ···················· | |
| Y | 3945 hCoV-19/England/ALDP-C3A3DB/2020\|EPI_ISL_731123\|20 | ···················· | ···················· | ···················· | ···················· | ···················· | ···················· | |
| Y | 3946 hCoV-19/England/ALDP-C3BA02/2020\|EPI_ISL_731124\|20 | ···················· | ···················· | ···················· | ···················· | ···················· | ···················· | |
| Y | 3947 hCoV-19/England/ALDP-C3CCB0/2020\|EPI_ISL_731125\|20 | ···················· | ···················· | ···················· | ···················· | ···················· | ···················· | |
| Y | 3948 hCoV-19/England/ALDP-C3A317/2020\|EPI_ISL_731131\|20 | ···················· | ·········T·········· | ···················· | ···················· | ···················· | ···················· | |
| Y | 3949 hCoV-19/England/ALDP-C3D04A/2020\|EPI_ISL_731132\|20 | ···················· | ···················· | ···················· | ···················· | ···················· | ···················· | |
| Y | 3950 hCoV-19/England/ALDP-C39EEA/2020\|EPI_ISL_731133\|20 | ···················· | ···················· | ···················· | ···················· | ···················· | ···················· | |
| Y | 3951 hCoV-19/England/ALDP-C39BF2/2020\|EPI_ISL_731135\|20 | ···················· | ···················· | ···················· | ···················· | ···················· | ···················· | |
| Y | 3952 hCoV-19/Wales/ALDP-C3D217/2020\|EPI_ISL_731138\|2020 | ···················· | ···················· | ···················· | ···················· | ···················· | ···················· | |
| Y | 3953 hCoV-19/England/ALDP-C3BC0C/2020\|EPI_ISL_731140\|20 | ···················· | ···················· | ···················· | ···················· | ···················· | ···················· | |
| Y | 3954 hCoV-19/England/ALDP-C39CA4/2020\|EPI_ISL_731141\|20 | ···················· | ···················· | ···················· | ···················· | ···················· | ···················· | |
| Y | 3955 hCoV-19/England/ALDP-C3C8DA/2020\|EPI_ISL_731142\|20 | ···················· | ···················· | ···················· | ···················· | ···················· | ···················· | |
| Y | 3956 hCoV-19/England/ALDP-C3CEE7/2020\|EPI_ISL_731144\|20 | ···················· | ···················· | ···················· | ···················· | ···················· | ···················· | |
| Y | 3957 hCoV-19/England/ALDP-C3C2EB/2020\|EPI_ISL_731145\|20 | ···················· | ···················· | ···················· | ···················· | ···················· | ···················· | |
| Y | 3958 hCoV-19/England/ALDP-C3A010/2020\|EPI_ISL_731147\|20 | ···················· | ···················· | ···················· | ···················· | ···················· | ···················· | |
| Y | 3959 hCoV-19/England/ALDP-C3C7BF/2020\|EPI_ISL_731149\|20 | ···················· | ···················· | ···················· | ···················· | ···················· | ···················· | |
| Y | 3960 hCoV-19/England/ALDP-C3D4D5/2020\|EPI_ISL_731152\|20 | ···················· | ···················· | ···················· | ···················· | ···················· | ···················· | |
| Y | 3961 hCoV-19/England/ALDP-C39ECC/2020\|EPI_ISL_731154\|20 | ···················· | ···················· | ···················· | ···················· | ···················· | ···················· | |
| Y | 3962 hCoV-19/England/ALDP-C3C791/2020\|EPI_ISL_731155\|20 | ···················· | ···················· | ···················· | ···················· | ···A················ | ···················· | |
| Y | 3963 hCoV-19/England/ALDP-C3BB96/2020\|EPI_ISL_731156\|20 | ···················· | ···················· | ···················· | ···················· | ···················· | ···················· | |
| Y | 3964 hCoV-19/England/ALDP-C3A405/2020\|EPI_ISL_731157\|20 | ···················· | ···················· | ···················· | ···················· | ···················· | ···················· | |
| Y | 3965 hCoV-19/England/ALDP-C3C97D/2020\|EPI_ISL_731159\|20 | ···················· | ···················· | ···················· | ···················· | ···················· | ···················· | |
| Y | 3966 hCoV-19/England/ALDP-C3A380/2020\|EPI_ISL_731160\|20 | ···················· | ···················· | ···················· | ···················· | ···················· | ···················· | |
| Y | 3967 hCoV-19/England/ALDP-C3A247/2020\|EPI_ISL_731161\|20 | ···················· | ···················· | ···················· | ···················· | ···················· | ···················· | |
| Y | 3968 hCoV-19/England/ALDP-C3B844/2020\|EPI_ISL_731163\|20 | ···················· | ···················· | ···················· | ···················· | ···················· | ···················· | |
| Y | 3969 hCoV-19/England/ALDP-C3CAD4/2020\|EPI_ISL_731164\|20 | ···················· | ···················· | ···················· | ···················· | ···················· | ···················· | |
| Y | 3970 hCoV-19/England/ALDP-C3BD90/2020\|EPI_ISL_731165\|20 | ···················· | ···················· | ···················· | ···················· | ···················· | ···················· | |
| Y | 3971 hCoV-19/England/ALDP-C3B9AB/2020\|EPI_ISL_731166\|20 | ···················· | ···················· | ···················· | ···················· | ···················· | ···················· | |
| Y | 3972 hCoV-19/England/ALDP-C3CC56/2020\|EPI_ISL_731168\|20 | ···················· | ···················· | ···················· | ···················· | ···················· | ···················· | |
| Y | 3973 hCoV-19/England/ALDP-C3BA20/2020\|EPI_ISL_731170\|20 | ···················· | ···················· | ···················· | ···················· | ···················· | ···················· | |
| Y | 3974 hCoV-19/England/ALDP-C3C649/2020\|EPI_ISL_731172\|20 | ···················· | ···················· | ···················· | ···················· | ···················· | ···A················ | |
| Y | 3975 hCoV-19/England/ALDP-C3DB85/2020\|EPI_ISL_731173\|20 | ···················· | ···················· | ···················· | ···················· | ···················· | ···················· | |
| Y | 3976 hCoV-19/England/ALDP-C3D0C2/2020\|EPI_ISL_731174\|20 | ···················· | ···················· | ···················· | ···················· | ···················· | ···················· | |
| Y | 3977 hCoV-19/England/ALDP-C3C48B/2020\|EPI_ISL_731175\|20 | ···················· | ···················· | ···················· | ···················· | ···················· | ···················· | |
| Y | 3978 hCoV-19/England/ALDP-C3D9B8/2020\|EPI_ISL_731176\|20 | ···················· | ···················· | ···················· | ···················· | ···················· | ···················· | |
| Y | 3979 hCoV-19/England/ALDP-C3C816/2020\|EPI_ISL_731177\|20 | ···················· | ···················· | ···················· | ···················· | ···················· | ···················· | |
| Y | 3980 hCoV-19/England/ALDP-C3CC83/2020\|EPI_ISL_731178\|20 | ···················· | ···················· | ···················· | ···················· | ···················· | ···················· | |
| Y | 3981 hCoV-19/England/ALDP-C3A283/2020\|EPI_ISL_731181\|20 | ···················· | ···················· | ···················· | ···················· | ···················· | ···················· | |
| Y | 3982 hCoV-19/England/ALDP-C3C46D/2020\|EPI_ISL_731182\|20 | ···················· | ···················· | ···················· | ···················· | ···················· | ···················· | |
| Y | 3983 hCoV-19/England/ALDP-C39C4A/2020\|EPI_ISL_731183\|20 | ···················· | ···················· | ···················· | ···················· | ···················· | ···················· | |
| Y | 3984 hCoV-19/England/ALDP-C3B71A/2020\|EPI_ISL_731184\|20 | ··················T· | ···················· | ···················· | ···················· | ···················· | ···················· | 1 |
| Y | 3985 hCoV-19/England/ALDP-C3BD63/2020\|EPI_ISL_731185\|20 | ···················· | ···················· | ···················· | ···················· | ···················· | ···················· | |
| Y | 3986 hCoV-19/England/ALDP-C3C500/2020\|EPI_ISL_731187\|20 | ···················· | ···················· | ···················· | ···················· | ···················· | ···················· | |
| Y | 3987 hCoV-19/England/ALDP-C3A292/2020\|EPI_ISL_731188\|20 | ···················· | ···················· | ···················· | ···················· | ···················· | ···················· | |
| Y | 3988 hCoV-19/England/ALDP-C3B747/2020\|EPI_ISL_731190\|20 | ···················· | ···················· | ···················· | ···················· | ···················· | ···················· | |
| Y | 3989 hCoV-19/England/ALDP-C3C272/2020\|EPI_ISL_731191\|20 | ···················· | ···················· | ···················· | ···················· | ···················· | ···················· | |
| Y | 3990 hCoV-19/England/ALDP-C3B941/2020\|EPI_ISL_731193\|20 | ···················· | ···················· | ···················· | ···················· | ···················· | ···················· | |
| Y | 3991 hCoV-19/England/ALDP-C3CA7A/2020\|EPI_ISL_731195\|20 | ···················· | ···················· | ···················· | ···················· | ···················· | ···················· | |
| Y | 3992 hCoV-19/England/ALDP-C3D97C/2020\|EPI_ISL_731198\|20 | ···················· | ···················· | ···················· | ···················· | ···················· | ···················· | |
| Y | 3993 hCoV-19/Ireland/D-NVRL-76IRL26419/2020\|EPI_ISL_732 | ···················· | ···················· | ···················· | ···················· | ···················· | ···················· | |
| Y | 3994 hCoV-19/Ireland/G-NVRL-76IRL33607/2020\|EPI_ISL_732 | ···················· | ···················· | ···················· | ···················· | ···················· | ···················· | |
| Y | 3995 hCoV-19/Ireland/KY-NVRL-76IRL33074/2020\|EPI_ISL_73 | ···················· | ···················· | ···················· | ···················· | ···················· | ···················· | |
| Y | 3996 hCoV-19/Ireland/SO-NVRL-76IRL31587/2020\|EPI_ISL_73 | ···················· | ···················· | ···················· | ···················· | ···················· | ···················· | |
| Y | 3997 hCoV-19/Ireland/WX-NVRL-76IRL29790/2020\|EPI_ISL_73 | ···················· | ···················· | ···················· | ···················· | ···················· | ···················· | |
| Y | 3998 hCoV-19/Ireland/CN-NVRL-76IRL32242/2020\|EPI_ISL_73 | ···················· | ···················· | ···················· | ···················· | ···················· | ···················· | |
| Y | 3999 hCoV-19/Ireland/CO-NVRL-76IRL32312/2020\|EPI_ISL_73 | ···················· | ···················· | ···················· | ···················· | ···················· | ···················· | |
| Y | 4000 hCoV-19/Ireland/CO-NVRL-76IRL32468/2020\|EPI_ISL_73 | ···················· | ···················· | ···················· | ···················· | ···················· | ···················· | |
| Y | 4001 hCoV-19/Ireland/CO-NVRL-76IRL32562/2020\|EPI_ISL_73 | ···················· | ···················· | ···················· | ···················· | ···················· | ···················· | |
| Y | 4002 hCoV-19/Ireland/G-NVRL-76IRL33308/2020\|EPI_ISL_732 | ···················· | ···················· | ···················· | ···················· | ···················· | ···················· | |
| Y | 4003 hCoV-19/Ireland/G-NVRL-76IRL33391/2020\|EPI_ISL_732 | ···················· | ···················· | ···················· | ···················· | ···················· | ···················· | |
| Y | 4004 hCoV-19/Ireland/G-NVRL-76IRL33539/2020\|EPI_ISL_732 | ···················· | ···················· | ···················· | ···················· | ···················· | ···················· | |
| Y | 4005 hCoV-19/Ireland/KE-NVRL-76IRL26522/2020\|EPI_ISL_73 | ···················· | ···················· | ···················· | ···················· | ···················· | ···················· | |
| Y | 4006 hCoV-19/Ireland/LH-NVRL-76IRL28214/2020\|EPI_ISL_73 | ···················· | ···················· | ···················· | ···················· | ···················· | ···················· | |
| Y | 4007 hCoV-19/Ireland/LK-NVRL-20ILR10653/2020\|EPI_ISL_73 | ···················· | ···················· | ···················· | ···················· | ···················· | ···················· | |
| Y | 4008 hCoV-19/Ireland/MH-NVRL-76IRL30430/2020\|EPI_ISL_73 | ···················· | ···················· | ···················· | ···················· | ···················· | ···················· | |
| Y | 4009 hCoV-19/Ireland/MH-NVRL-76IRL30440/2020\|EPI_ISL_73 | ···················· | ···················· | ···················· | ···················· | ···················· | ···················· | |
| Y | 4010 hCoV-19/Ireland/MN-NVRL-76IRL32789/2020\|EPI_ISL_73 | ···················· | ···················· | ···················· | ···················· | ···················· | ···················· | |
| Y | 4011 hCoV-19/Ireland/SO-NVRL-76IRL31413/2020\|EPI_ISL_73 | ···················· | ···················· | ···················· | ···················· | ···················· | ···················· | |
| Y | 4012 hCoV-19/Ireland/SO-NVRL-76IRL31614/2020\|EPI_ISL_73 | ···················· | ···················· | ···················· | ···················· | ···················· | ···················· | |
| Y | 4013 hCoV-19/Ireland/CO-NVRL-76IRL32367/2020\|EPI_ISL_73 | ···················· | ···················· | ···················· | ···················· | ···················· | ···················· | |
| Y | 4014 hCoV-19/Ireland/D-NVRL-76IRL32023/2020\|EPI_ISL_732 | ···················· | ···················· | ···················· | ···················· | ···················· | ···················· | |
| Y | 4015 hCoV-19/Ireland/D-NVRL-76IRL33884/2020\|EPI_ISL_732 | ···················· | ···················· | ···················· | ···················· | ···················· | ···················· | |
| Y | 4016 hCoV-19/Ireland/KE-NVRL-76IRL26703/2020\|EPI_ISL_73 | ···················· | ···················· | ···················· | ···················· | ···················· | ···················· | |
| Y | 4017 hCoV-19/Ireland/LH-NVRL-20ILR08017/2020\|EPI_ISL_73 | ···················· | ···················· | ···················· | ···················· | ···················· | ···················· | |
| Y | 4018 hCoV-19/Ireland/D-NVRL-76IRL32009/2020\|EPI_ISL_732 | ···················· | ···················· | ···················· | ···················· | ···················· | ···················· | |
| Y | 4020 hCoV-19/Ireland/LH-NVRL-76IRL28215/2020\|EPI_ISL_73 | ···················· | ···················· | ···················· | ···················· | ···················· | ···················· | |

| | | | | | | | |
|---|---|---|---|---|---|---|---|
| Y | 4021 hCoV-19/Ireland/LH-NVRL-76IRL28298/2020\|EPI_ISL_73 | ···················· | ···················· | ···················· | ···················· | ···················· | ···················· |
| Y | 4022 hCoV-19/Ireland/MH-NVRL-76IRL26873/2020\|EPI_ISL_73 | ···················· | ···················· | ···················· | ···················· | ···················· | ···················· |
| Y | 4023 hCoV-19/Ireland/MH-NVRL-76IRL32072/2020\|EPI_ISL_73 | ···················· | ···················· | ···················· | ···················· | ···················· | ···················· |
| Y | 4024 hCoV-19/Ireland/OY-NVRL-76IRL30545/2020\|EPI_ISL_73 | ···················· | ···················· | ···················· | ···················· | ···················· | ···················· |
| Y | 4025 hCoV-19/Ireland/D-NVRL-76IRL28438/2020\|EPI_ISL_732 | ···················· | ···················· | ···················· | ···················· | ···················· | ···················· |
| Y | 4026 hCoV-19/Ireland/D-NVRL-76IRL31920/2020\|EPI_ISL_732 | ···················· | ····A··············· | ···················· | ···················· | ···················· | ···················· |
| Y | 4027 hCoV-19/Ireland/KK-NVRL-76IRL28885/2020\|EPI_ISL_73 | ···················· | ···················· | ···················· | ···················· | ···················· | ···················· |
| Y | 4028 hCoV-19/Ireland/KE-NVRL-78IRL48764/2020\|EPI_ISL_73 | ···················· | ···················· | ···················· | ···················· | ···················· | ···················· |
| Y | 4029 hCoV-19/Ireland/MH-NVRL-76IRL27100/2020\|EPI_ISL_73 | ···················· | ···················· | ···················· | ···················· | ···················· | ···················· |
| Y | 4030 hCoV-19/Ireland/MO-NVRL-79IRL03919/2020\|EPI_ISL_73 | ···················· | ···················· | ···················· | ···················· | ···················· | ···················· |
| Y | 4031 hCoV-19/Ireland/MO-NVRL-76IRL31264/2020\|EPI_ISL_73 | ···················· | ···················· | ···················· | ···················· | ···················· | ···················· |
| Y | 4032 hCoV-19/Ireland/D-NVRL-78IRL92230/2020\|EPI_ISL_732 | ···················· | ···················· | ···················· | ···················· | ···················· | ···················· |
| Y | 4033 hCoV-19/Ireland/D-NVRL-20IRL93293/2020\|EPI_ISL_732 | ···················· | ···················· | ···················· | ···················· | ···················· | ···················· |
| Y | 4034 hCoV-19/Ireland/D-NVRL-78IRL92235/2020\|EPI_ISL_732 | ···················· | ···················· | ···················· | ···················· | ···················· | ···················· |
| Y | 4035 hCoV-19/England/ALDP-C22F3C/2020\|EPI_ISL_733701\|20 | ···················· | ···················· | ···················· | ···················· | ···················· | ···················· |
| Y | 4036 hCoV-19/England/CAMC-C2CEBD/2020\|EPI_ISL_733702\|20 | ···················· | ···················· | ···················· | ···················· | ···················· | ···················· |
| Y | 4037 hCoV-19/England/ALDP-C22ED5/2020\|EPI_ISL_733703\|20 | ···················· | ···················· | ···················· | ···················· | ···················· | ···················· |
| Y | 4038 hCoV-19/England/ALDP-C230FC/2020\|EPI_ISL_733710\|20 | ···················· | ···················· | ···················· | ···················· | ···················· | ···················· |
| Y | 4039 hCoV-19/England/MILK-C2F9F5/2020\|EPI_ISL_733711\|20 | ···················· | ···················· | ···················· | ···················· | ···················· | ···················· |
| Y | 4040 hCoV-19/England/ALDP-C230A1/2020\|EPI_ISL_733712\|20 | ···················· | ···················· | ···················· | ···················· | ···················· | ···················· |
| Y | 4041 hCoV-19/England/CAMC-C268E2/2020\|EPI_ISL_733715\|20 | ···················· | ···················· | ···················· | ···················· | ···················· | ···················· |
| Y | 4042 hCoV-19/England/CAMC-C2CFBA/2020\|EPI_ISL_733716\|20 | ···················· | ···················· | ···················· | ···················· | ···················· | ···················· |
| Y | 4043 hCoV-19/England/CAMC-C268D3/2020\|EPI_ISL_733722\|20 | ···················· | ···················· | ···················· | ···················· | ···················· | ···················· |
| Y | 4044 hCoV-19/England/ALDP-C230ED/2020\|EPI_ISL_733725\|20 | ···················· | ···················· | ···················· | ···················· | ···················· | ···················· |
| Y | 4045 hCoV-19/England/CAMC-C2C943/2020\|EPI_ISL_733726\|20 | ···················· | ···················· | ···················· | ···················· | ···················· | ···················· |
| Y | 4046 hCoV-19/England/CAMC-C2CECC/2020\|EPI_ISL_733730\|20 | ···················· | ···················· | ···················· | ···················· | ···················· | ···················· |
| Y | 4047 hCoV-19/England/ALDP-C22EF3/2020\|EPI_ISL_733731\|20 | ···················· | ···················· | ···················· | ···················· | ···················· | ···················· |
| Y | 4048 hCoV-19/England/CAMC-C2D3F9/2020\|EPI_ISL_733732\|20 | ···················· | ···················· | ···················· | ···················· | ···················· | ···················· |
| Y | 4049 hCoV-19/England/CAMC-C26439/2020\|EPI_ISL_733733\|20 | ···················· | ·A·················· | ···················· | ···················· | ···················· | ···A················ |
| Y | 4050 hCoV-19/England/ALDP-C23232/2020\|EPI_ISL_733734\|20 | ···················· | ···················· | ···················· | ···················· | ···················· | ···················· |
| Y | 4051 hCoV-19/England/MILK-C2FC56/2020\|EPI_ISL_733735\|20 | ···················· | ···················· | ···················· | ···················· | ···················· | ···················· |
| Y | 4052 hCoV-19/England/ALDP-C23241/2020\|EPI_ISL_733737\|20 | ···················· | ···················· | ···················· | ···················· | ···················· | ···················· |
| Y | 4053 hCoV-19/England/MILK-C2FB86/2020\|EPI_ISL_733739\|20 | ···················· | ···················· | ···················· | ···················· | ···················· | ···················· |
| Y | 4054 hCoV-19/England/CAMC-C2D317/2020\|EPI_ISL_733741\|20 | ···················· | ···················· | ···················· | ···················· | ···················· | ···················· |
| Y | 4055 hCoV-19/England/ALDP-C233D5/2020\|EPI_ISL_733742\|20 | ···················· | ···················· | ···················· | ···················· | ···················· | ···A················ |
| Y | 4056 hCoV-19/England/MILK-C2F95F/2020\|EPI_ISL_733746\|20 | ···················· | ···················· | ···················· | ···················· | ···················· | ···················· |
| Y | 4057 hCoV-19/England/MILK-C2FC38/2020\|EPI_ISL_733749\|20 | ···················· | ···················· | ···················· | ···················· | ···················· | ···················· |
| Y | 4058 hCoV-19/England/ALDP-C233F3/2020\|EPI_ISL_733754\|20 | ···················· | ···················· | ···················· | ···················· | ···················· | ···················· |
| Y | 4059 hCoV-19/England/ALDP-C23171/2020\|EPI_ISL_733755\|20 | ···················· | ···················· | ···················· | ···················· | ···················· | ···················· |
| Y | 4060 hCoV-19/England/MILK-C305C0/2020\|EPI_ISL_733757\|20 | ···················· | ···················· | ···················· | ···················· | ···················· | ···················· |
| Y | 4061 hCoV-19/England/CAMC-C2CF9C/2020\|EPI_ISL_733758\|20 | ···················· | ···················· | ···················· | ···················· | ···················· | ···················· |
| Y | 4062 hCoV-19/England/ALDP-C23047/2020\|EPI_ISL_733759\|20 | ···················· | ···················· | ···················· | ···················· | ···················· | ···················· |
| Y | 4063 hCoV-19/England/MILK-C30575/2020\|EPI_ISL_733760\|20 | ···················· | ···················· | ···················· | ···················· | ···················· | ···················· |
| Y | 4064 hCoV-19/England/MILK-C2F97D/2020\|EPI_ISL_733764\|20 | ···················· | ···················· | ···················· | ···················· | ···················· | ···················· |
| Y | 4065 hCoV-19/England/CAMC-C2CAAA/2020\|EPI_ISL_733765\|20 | ···················· | ···················· | ···················· | ···················· | ···················· | ···················· |
| Y | 4066 hCoV-19/England/ALDP-C23C52/2020\|EPI_ISL_733766\|20 | ···················· | ···················· | ···················· | ···················· | ···················· | ···················· |
| Y | 4067 hCoV-19/England/MILK-C2F6D0/2020\|EPI_ISL_733768\|20 | ···················· | ···················· | ···················· | ···················· | ···················· | ···················· |
| Y | 4068 hCoV-19/England/ALDP-C2326F/2020\|EPI_ISL_733769\|20 | ···················· | ···················· | ···················· | ···················· | ···················· | ···A················ |
| Y | 4069 hCoV-19/England/MILK-C2FC74/2020\|EPI_ISL_733771\|20 | ···················· | ···················· | ···················· | ···················· | ···················· | ···················· |
| Y | 4070 hCoV-19/England/ALDP-C23CDA/2020\|EPI_ISL_733777\|20 | ···················· | ···················· | ···················· | ···················· | ······T············· | ···················· |
| Y | 4071 hCoV-19/England/CAMC-C26150/2020\|EPI_ISL_733778\|20 | ···················· | ···················· | ···················· | ···················· | ···················· | ···················· |
| Y | 4072 hCoV-19/England/ALDP-C23126/2020\|EPI_ISL_733783\|20 | ···················· | ···················· | ···················· | ···················· | ···················· | ···················· |
| Y | 4073 hCoV-19/England/CAMC-C2CFD8/2020\|EPI_ISL_733784\|20 | ···················· | ···················· | ···················· | ···················· | ···················· | ···················· |
| Y | 4074 hCoV-19/England/CAMC-C2CEDB/2020\|EPI_ISL_733786\|20 | ···················· | ···················· | ···················· | ···················· | ···················· | ···················· |
| Y | 4075 hCoV-19/England/CAMC-C2CF41/2020\|EPI_ISL_733787\|20 | ···················· | ·A·················· | ···················· | ···················· | ···················· | ···A················ |
| Y | 4076 hCoV-19/England/CAMC-C2D353/2020\|EPI_ISL_733793\|20 | ···················· | ···················· | ···················· | ···················· | ···················· | ···················· |
| Y | 4077 hCoV-19/England/ALDP-C23083/2020\|EPI_ISL_733794\|20 | ···················· | ···················· | ···················· | ···················· | ···················· | ···················· |
| Y | 4078 hCoV-19/England/ALDP-C23162/2020\|EPI_ISL_733795\|20 | ···················· | ·A·················· | ···················· | ···················· | ···················· | ···················· |
| Y | 4079 hCoV-19/England/MILK-C2F9D7/2020\|EPI_ISL_733796\|20 | ···················· | ···················· | ···················· | ···················· | ···················· | ···················· |
| Y | 4080 hCoV-19/England/ALDP-C22EB7/2020\|EPI_ISL_733797\|20 | ···················· | ···················· | ···················· | ···················· | ···················· | ···················· |
| Y | 4081 hCoV-19/England/MILK-C2FB59/2020\|EPI_ISL_733798\|20 | ···················· | ···················· | ···················· | ···················· | ···················· | ···················· |
| Y | 4082 hCoV-19/Finland/13LS14AE1/2020\|EPI_ISL_737220\|2020 | ···················· | ·A·················· | ···················· | ···················· | ···················· | ···A················ |
| Y | 4083 hCoV-19/Finland/13LS14AH1/2020\|EPI_ISL_737221\|2020 | ···················· | ···················· | ···················· | ···················· | ···················· | ···················· |
| Y | 4084 hCoV-19/Finland/13LS15BD5/2020\|EPI_ISL_737222\|2020 | ···················· | ·A·················· | ···················· | ···················· | ···················· | ···A················ |
| Y | 4085 hCoV-19/Finland/13LS16AH3/2020\|EPI_ISL_737223\|2020 | ···················· | ·A·················· | ···················· | ···················· | ···················· | ···A················ |
| Y | 4086 hCoV-19/Finland/13LS17G8/2020\|EPI_ISL_737224\|2020- | ···················· | ·A·················· | ···················· | ···················· | ···················· | ···A················ |
| Y | 4087 hCoV-19/Finland/13LS20AD9/2020\|EPI_ISL_737225\|2020 | ···················· | ···················· | ···················· | ···················· | ···················· | ···················· |
| Y | 4088 hCoV-19/Finland/13LS3AB7/2020\|EPI_ISL_737226\|2020- | ···················· | ·A·················· | ···················· | ···················· | ···················· | ···A················ |
| Y | 4089 hCoV-19/Finland/13LS3AD5/2020\|EPI_ISL_737227\|2020- | ···················· | ···················· | ···················· | ···················· | ···················· | ···················· |
| Y | 4090 hCoV-19/Finland/13LS4AG7/2020\|EPI_ISL_737228\|2020- | ···················· | ···················· | ···················· | ···················· | ···················· | ···················· |
| Y | 4091 hCoV-19/Finland/13LS5AD3/2020\|EPI_ISL_737229\|2020- | ···················· | ·A·················· | ···················· | ···················· | ···················· | ···A················ |
| Y | 4092 hCoV-19/Finland/13LS6AG7/2020\|EPI_ISL_737230\|2020- | ···················· | ···················· | ···················· | ···················· | ···················· | ···················· |
| Y | 4093 hCoV-19/Finland/13LS7AH2/2020\|EPI_ISL_737231\|2020- | ···················· | ···················· | ···················· | ···················· | ···················· | ···················· |
| Y | 4094 hCoV-19/Finland/13SS15B9/2020\|EPI_ISL_737234\|2020- | ···················· | ···················· | ···················· | ···················· | ···················· | ···················· |
| Y | 4095 hCoV-19/Finland/13SS5B11/2020\|EPI_ISL_737235\|2020- | ···················· | ···················· | ···················· | ···················· | ···················· | ···················· |
| Y | 4096 hCoV-19/USA/CA-LACPHL-AF00025/2020\|EPI_ISL_737260\| | ···················· | ···················· | ···················· | ···················· | ···················· | ···················· |
| Y | 4097 hCoV-19/England/NORW-F7084/2020\|EPI_ISL_725323\|202 | ···················· | ···················· | ···················· | ···················· | ···················· | ···················· |
| Y | 4098 hCoV-19/England/NORW-F7093/2020\|EPI_ISL_725324\|202 | ···················· | ···················· | ···················· | ···················· | ···················· | ···················· |
| Y | 4099 hCoV-19/England/NORW-F70DF/2020\|EPI_ISL_725326\|202 | ···················· | ···················· | ···················· | ···················· | ···················· | ···················· |
| Y | 4100 hCoV-19/England/NORW-F70EE/2020\|EPI_ISL_725327\|202 | ···················· | ···················· | ···················· | ···················· | ···················· | ···················· |

| | | | | | | | | |
|---|---|---|---|---|---|---|---|---|
| Y | 4101 | hCoV-19/England/NORW-F70FD/2020\|EPI_ISL_725328\|202 | ···················· | ···················· | ···················· | ···················· | ···················· | ···················· |
| Y | 4102 | hCoV-19/England/NORW-F7109/2020\|EPI_ISL_725329\|202 | ···················· | ···················· | ···················· | ···················· | ···················· | ···················· |
| Y | 4103 | hCoV-19/England/NORW-F7127/2020\|EPI_ISL_725331\|202 | ···················· | ···················· | ···················· | ···················· | ···················· | ···················· |
| Y | 4104 | hCoV-19/England/NORW-F7497/2020\|EPI_ISL_725332\|202 | ···················· | ···················· | ···················· | ···················· | ···················· | ···················· |
| Y | 4106 | hCoV-19/England/NORW-F75EF/2020\|EPI_ISL_725336\|202 | ···················· | ···················· | ···················· | ···················· | ···················· | ···················· |
| Y | 4107 | hCoV-19/England/NORW-F7655/2020\|EPI_ISL_725338\|202 | ···················· | ···················· | ···················· | ···················· | ···················· | ···················· |
| Y | 4108 | hCoV-19/England/NORW-F76DD/2020\|EPI_ISL_725339\|202 | ···················· | ···················· | ···················· | ···················· | ···················· | ···················· |
| Y | 4110 | hCoV-19/England/NORW-F7734/2020\|EPI_ISL_725342\|202 | ···················· | ···················· | ···················· | ···················· | ···················· | ···················· |
| Y | 4111 | hCoV-19/England/NORW-F8593/2020\|EPI_ISL_725343\|202 | ···················· | ·A·················· | ···················· | ···················· | ···················· | ··A················· |
| Y | 4112 | hCoV-19/England/NORW-F85A2/2020\|EPI_ISL_725344\|202 | ···················· | ·A·················· | ···················· | ···················· | ···················· | ··A················· |
| Y | 4113 | hCoV-19/England/NORW-F8627/2020\|EPI_ISL_725348\|202 | ···················· | ···················· | ···················· | ···················· | ···················· | ···················· |
| Y | 4114 | hCoV-19/England/NORW-F8645/2020\|EPI_ISL_725349\|202 | ···················· | ···················· | ···················· | ···················· | ···················· | ···················· |
| Y | 4115 | hCoV-19/England/NORW-F86BE/2020\|EPI_ISL_725354\|202 | ···················· | ···················· | ···················· | ···················· | ···················· | ···················· |
| Y | 4116 | hCoV-19/England/NORW-F8733/2020\|EPI_ISL_725359\|202 | ···················· | ···················· | ···················· | ···················· | ···················· | ···················· |
| Y | 4117 | hCoV-19/England/NORW-F878E/2020\|EPI_ISL_725362\|202 | ···················· | ···················· | ···················· | ···················· | ···················· | ···················· |
| Y | 4118 | hCoV-19/England/NORW-F879D/2020\|EPI_ISL_725363\|202 | ···················· | ···················· | ···················· | ···················· | ···················· | ···················· |
| Y | 4119 | hCoV-19/England/NORW-F87BB/2020\|EPI_ISL_725365\|202 | ···················· | ···················· | ···················· | ···················· | ···················· | ···················· |
| Y | 4120 | hCoV-19/England/NORW-F887C/2020\|EPI_ISL_725368\|202 | ···················· | ···················· | ···················· | ···················· | ···················· | ···················· |
| Y | 4121 | hCoV-19/England/NORW-F8C52/2020\|EPI_ISL_725371\|202 | ···················· | ···················· | ···················· | ···················· | ···················· | ···················· |
| Y | 4122 | hCoV-19/England/NORW-F8CBC/2020\|EPI_ISL_725374\|202 | ···················· | ·A·················· | ···················· | ···················· | ···················· | ··A················· |
| Y | 4123 | hCoV-19/England/NORW-F8DAA/2020\|EPI_ISL_725376\|202 | ···················· | ···················· | ···················· | ···················· | ···················· | ···················· |
| Y | 4124 | hCoV-19/England/NORW-F8DB9/2020\|EPI_ISL_725377\|202 | ···················· | ···················· | ···················· | ···················· | ···················· | ···················· |
| Y | 4125 | hCoV-19/England/NORW-F8EB6/2020\|EPI_ISL_725379\|202 | ···················· | ···················· | ···················· | ···················· | ···················· | ···················· |
| Y | 4126 | hCoV-19/England/NORW-F8FD1/2020\|EPI_ISL_725381\|202 | ···················· | ···················· | ···················· | ···················· | ···················· | ···················· |
| Y | 4127 | hCoV-19/England/NORW-F9468/2020\|EPI_ISL_725384\|202 | ···················· | ···················· | ···················· | ···················· | ···················· | ···················· |
| Y | 4128 | hCoV-19/England/NORW-F9574/2020\|EPI_ISL_725386\|202 | ···················· | ···················· | ···················· | ···················· | ···················· | ···················· |
| Y | 4129 | hCoV-19/England/NORW-F9635/2020\|EPI_ISL_725389\|202 | ···················· | ···················· | ···················· | ···················· | ···················· | ···················· |
| Y | 4130 | hCoV-19/England/NORW-F96BD/2020\|EPI_ISL_725391\|202 | ···················· | ···················· | ···················· | ···················· | ···················· | ···················· |
| Y | 4131 | hCoV-19/England/NORW-F976F/2020\|EPI_ISL_725393\|202 | ···················· | ···················· | ···················· | ···················· | ···················· | ···················· |
| Y | 4132 | hCoV-19/England/NORW-F97D8/2020\|EPI_ISL_725396\|202 | ···················· | ···················· | ···················· | ···················· | ···················· | ···················· |
| Y | 4133 | hCoV-19/England/NORW-F9802/2020\|EPI_ISL_725398\|202 | ···················· | ···················· | ···················· | ···················· | ···················· | ···················· |
| Y | 4134 | hCoV-19/England/NORW-F985D/2020\|EPI_ISL_725399\|202 | ···················· | ···················· | ···················· | ···················· | ···················· | ···················· |
| Y | 4135 | hCoV-19/Wales/PHWC-4965EE/2020\|EPI_ISL_726607\|2020 | ···················· | ···················· | ···················· | ···················· | ···················· | ···················· |
| Y | 4136 | hCoV-19/Wales/PHWC-4965FD/2020\|EPI_ISL_726608\|2020 | ···················· | ···················· | ···················· | ···················· | ···················· | ···················· |
| Y | 4137 | hCoV-19/Wales/PHWC-496609/2020\|EPI_ISL_726609\|2020 | ···················· | ···················· | ···················· | ···················· | ···················· | ···················· |
| Y | 4138 | hCoV-19/Wales/PHWC-496618/2020\|EPI_ISL_726610\|2020 | ···················· | ···················· | ···················· | ···················· | ···················· | ···················· |
| Y | 4140 | hCoV-19/Wales/PHWC-496690/2020\|EPI_ISL_726618\|2020 | ···················· | ···················· | ···················· | ···················· | ···················· | ···················· |
| Y | 4141 | hCoV-19/Wales/PHWC-4966AF/2020\|EPI_ISL_726619\|2020 | ···················· | ···················· | ···················· | ···················· | ···················· | ···················· |
| Y | 4142 | hCoV-19/Wales/PHWC-4966BE/2020\|EPI_ISL_726620\|2020 | ···················· | ···················· | ···················· | ···················· | ···················· | ···················· |
| Y | 4144 | hCoV-19/Wales/PHWC-4966DC/2020\|EPI_ISL_726622\|2020 | ···················· | ···················· | ···················· | ···················· | ···················· | ···················· |
| Y | 4145 | hCoV-19/Wales/PHWC-4966EB/2020\|EPI_ISL_726623\|2020 | ···················· | ···················· | ···················· | ···················· | ···················· | ···················· |
| Y | 4146 | hCoV-19/Wales/PHWC-4966FA/2020\|EPI_ISL_726624\|2020 | ···················· | ···················· | ···················· | ···················· | ···················· | ···················· |
| Y | 4147 | hCoV-19/Wales/PHWC-496706/2020\|EPI_ISL_726625\|2020 | ···················· | ···················· | ···················· | ···················· | ···················· | ···················· |
| Y | 4148 | hCoV-19/Wales/PHWC-496715/2020\|EPI_ISL_726626\|2020 | ···················· | ···················· | ···················· | ···················· | ···················· | ···················· |
| Y | 4149 | hCoV-19/Wales/PHWC-496724/2020\|EPI_ISL_726627\|2020 | ···················· | ···················· | ···················· | ···················· | ···················· | ···················· |
| Y | 4150 | hCoV-19/Wales/PHWC-496733/2020\|EPI_ISL_726628\|2020 | ···················· | ···················· | ···················· | ···················· | ···················· | ···················· |
| Y | 4151 | hCoV-19/Wales/PHWC-4968A9/2020\|EPI_ISL_726650\|2020 | ···················· | ···················· | ···················· | ···················· | ···················· | ···················· |
| Y | 4153 | hCoV-19/Wales/PHWC-4968D6/2020\|EPI_ISL_726653\|2020 | ···················· | ···················· | ···················· | ···················· | ···················· | ···················· |
| Y | 4154 | hCoV-19/Wales/PHWC-4968E5/2020\|EPI_ISL_726654\|2020 | ···················· | ···················· | ···················· | ···················· | ···················· | ···················· |
| Y | 4156 | hCoV-19/Wales/PHWC-496900/2020\|EPI_ISL_726656\|2020 | ···················· | ···················· | ···················· | ···················· | ···················· | ···················· |
| Y | 4161 | hCoV-19/Wales/PHWC-496997/2020\|EPI_ISL_726664\|2020 | ···················· | ···················· | ···················· | ···················· | ···················· | ···················· |
| Y | 4167 | hCoV-19/Wales/PHWC-496A1C/2020\|EPI_ISL_726672\|2020 | ···················· | ···················· | ···················· | ···················· | ···················· | ···················· |
| Y | 4169 | hCoV-19/Wales/PHWC-496A3A/2020\|EPI_ISL_726674\|2020 | ···················· | ···················· | ···················· | ···················· | ···················· | ···················· |
| Y | 4171 | hCoV-19/Wales/PHWC-496A76/2020\|EPI_ISL_726678\|2020 | ···················· | ···················· | ···················· | ···················· | ···················· | ···················· |
| Y | 4172 | hCoV-19/Wales/PHWC-496A85/2020\|EPI_ISL_726679\|2020 | ···················· | ···················· | ···················· | ···················· | ···················· | ···················· |
| Y | 4176 | hCoV-19/Wales/PHWC-496AD0/2020\|EPI_ISL_726684\|2020 | ···················· | ···················· | ···················· | ···················· | ···················· | ···················· |
| Y | 4179 | hCoV-19/Wales/PHWC-496B0A/2020\|EPI_ISL_726687\|2020 | ···················· | ···················· | ···················· | ···················· | ···················· | ···················· |
| Y | 4180 | hCoV-19/Wales/PHWC-496B19/2020\|EPI_ISL_726688\|2020 | ···················· | ···················· | ···················· | ···················· | ···················· | ···················· |
| Y | 4181 | hCoV-19/Wales/PHWC-496B37/2020\|EPI_ISL_726690\|2020 | ···················· | ···················· | ···················· | ···················· | ···················· | ···················· |
| Y | 4183 | hCoV-19/Wales/PHWC-496B55/2020\|EPI_ISL_726692\|2020 | ···················· | ···················· | ···················· | ···················· | ···················· | ···················· |
| Y | 4184 | hCoV-19/Wales/PHWC-496B64/2020\|EPI_ISL_726693\|2020 | ···················· | ···················· | ···················· | ···················· | ···················· | ···················· |
| Y | 4185 | hCoV-19/Wales/PHWC-496B73/2020\|EPI_ISL_726694\|2020 | ···················· | ···················· | ···················· | ···················· | ···················· | ···················· |
| Y | 4187 | hCoV-19/Wales/PHWC-496BBF/2020\|EPI_ISL_726697\|2020 | ···················· | ···················· | ···················· | ···················· | ···················· | ···················· |
| Y | 4189 | hCoV-19/England/SHEF-10D032B/2020\|EPI_ISL_727938\|2 | ···················· | ···················· | ···················· | ···················· | ···················· | ···················· |
| Y | 4190 | hCoV-19/England/SHEF-10D0CE4/2020\|EPI_ISL_727939\|2 | ···················· | ···················· | ···················· | ···················· | ···················· | ···················· |
| Y | 4191 | hCoV-19/England/SHEF-10D0DC3/2020\|EPI_ISL_727940\|2 | ···················· | ···················· | ···················· | ···················· | ···················· | ···················· |
| Y | 4192 | hCoV-19/England/SHEF-10D1D59/2020\|EPI_ISL_727950\|2 | ···················· | ···················· | ···················· | ···················· | ···················· | ···················· |
| Y | 4193 | hCoV-19/England/SHEF-10D2945/2020\|EPI_ISL_727959\|2 | ···················· | ···················· | ···················· | ···················· | ···················· | ···················· |
| Y | 4194 | hCoV-19/England/SHEF-10D6D54/2020\|EPI_ISL_727982\|2 | ···················· | ···················· | ···················· | ···················· | ···················· | ···················· |
| Y | 4195 | hCoV-19/England/SHEF-10D6F8B/2020\|EPI_ISL_727983\|2 | ···················· | ···················· | ···················· | ···················· | ···················· | ···················· |
| Y | 4196 | hCoV-19/England/SHEF-10D703C/2020\|EPI_ISL_727984\|2 | ···················· | ···················· | ···················· | ···················· | ·····T·············· | ···················· |
| Y | 4197 | hCoV-19/England/SHEF-10D73E8/2020\|EPI_ISL_727985\|2 | ···················· | ···················· | ···················· | ···················· | ···················· | ···················· |
| Y | 4198 | hCoV-19/England/SHEF-10D7C56/2020\|EPI_ISL_727986\|2 | ···················· | ···················· | ···················· | ···················· | ···················· | ···················· |
| Y | 4199 | hCoV-19/England/SHEF-10D83E7/2020\|EPI_ISL_727987\|2 | ···················· | ···················· | ···················· | ···················· | ···················· | ···················· |
| Y | 4200 | hCoV-19/England/SHEF-10D8745/2020\|EPI_ISL_727988\|2 | ···················· | ···················· | ···················· | ···················· | ···················· | ···················· |
| Y | 4201 | hCoV-19/England/SHEF-C82E0/2020\|EPI_ISL_727993\|202 | ···················· | ···················· | ···················· | ···················· | ···················· | ···················· |
| Y | 4202 | hCoV-19/England/SHEF-CA2A2/2020\|EPI_ISL_727994\|202 | ···················· | ···················· | ···················· | ···················· | ···················· | ···················· |
| Y | 4203 | hCoV-19/England/SHEF-D27E4/2020\|EPI_ISL_727997\|202 | ···················· | ···················· | ···················· | ···················· | ···················· | ···················· |
| Y | 4204 | hCoV-19/Japan/FHU-CS27-0221/2020\|EPI_ISL_727998\|20 | ···················· | ···················· | ···················· | ···················· | ···················· | ···················· |
| Y | 4205 | hCoV-19/Japan/FHU-CS29-0221/2020\|EPI_ISL_727999\|20 | ···················· | ···················· | ···················· | ···················· | ···················· | ···················· |

| | | | | | | | | |
|---|---|---|---|---|---|---|---|---|
| Y | 4206 | hCoV-19/Ireland/LH-NVRL-20ILR11476/2020\|EPI_ISL_73 | ···················· | ···················· | ···················· | ···················· | ···················· | ···················· |
| Y | 4207 | hCoV-19/Ireland/LH-NVRL-20ILR11477/2020\|EPI_ISL_73 | ···················· | ···················· | ···················· | ···················· | ···················· | ···················· |
| Y | 4208 | hCoV-19/Ireland/LH-NVRL-20ILR11478/2020\|EPI_ISL_73 | ···················· | ···················· | ···················· | ···················· | ···················· | ···················· |
| Y | 4209 | hCoV-19/Ireland/LH-NVRL-20ILR11479/2020\|EPI_ISL_73 | ···················· | ···················· | ···················· | ···················· | ···················· | ···················· |
| Y | 4210 | hCoV-19/Ireland/LH-NVRL-20ILR11480/2020\|EPI_ISL_73 | ···················· | ···················· | ···················· | ···················· | ···················· | ···················· |
| Y | 4211 | hCoV-19/Ireland/LH-NVRL-20ILR11483/2020\|EPI_ISL_73 | ···················· | ···················· | ···················· | ···················· | ···················· | ···················· |
| Y | 4212 | hCoV-19/Ireland/LH-NVRL-20ILR11484/2020\|EPI_ISL_73 | ···················· | ···················· | ···················· | ···················· | ···················· | ···················· |
| Y | 4213 | hCoV-19/Ireland/LH-NVRL-20ILR11485/2020\|EPI_ISL_73 | ···················· | ···················· | ···················· | ···················· | ···················· | ···················· |
| Y | 4214 | hCoV-19/Ireland/LH-NVRL-20ILR11487/2020\|EPI_ISL_73 | ···················· | ···················· | ···················· | ···················· | ···················· | ···················· |
| Y | 4215 | hCoV-19/Ireland/LH-NVRL-20ILR11488/2020\|EPI_ISL_73 | ···················· | ···················· | ···················· | ···················· | ···················· | ···················· |
| Y | 4216 | hCoV-19/Ireland/LH-NVRL-20ILR11489/2020\|EPI_ISL_73 | ···················· | ···················· | ···················· | ···················· | ···················· | ···················· |
| Y | 4217 | hCoV-19/Ireland/LH-NVRL-20ILR11490/2020\|EPI_ISL_73 | ···················· | ···················· | ···················· | ···················· | ···················· | ···················· |
| Y | 4218 | hCoV-19/Ireland/LH-NVRL-20ILR11491/2020\|EPI_ISL_73 | ···················· | ···················· | ···················· | ···················· | ···················· | ···················· |
| Y | 4219 | hCoV-19/Ireland/LH-NVRL-20ILR11492/2020\|EPI_ISL_73 | ···················· | ···················· | ···················· | ···················· | ···················· | ···················· |
| Y | 4220 | hCoV-19/Ireland/CE-NVRL-74IRL52231/2020\|EPI_ISL_73 | ···················· | ···················· | ···················· | ···················· | ···················· | ···················· |
| Y | 4221 | hCoV-19/Ireland/CN-NVRL-76IRL32236/2020\|EPI_ISL_73 | ···················· | ···················· | ···················· | ···················· | ···················· | ···················· |
| Y | 4222 | hCoV-19/Ireland/CO-NVRL-76IRL32511/2020\|EPI_ISL_73 | ···················· | ···················· | ···················· | ···················· | ···················· | ···················· |
| Y | 4223 | hCoV-19/Ireland/G-NVRL-76IRL33626/2020\|EPI_ISL_732 | ···················· | ···················· | ···················· | ···················· | ···················· | ···················· |
| Y | 4224 | hCoV-19/Ireland/KE-NVRL-76IRL26610/2020\|EPI_ISL_73 | ···················· | ···················· | ···················· | ···················· | ···················· | ···················· |
| Y | 4225 | hCoV-19/Ireland/KE-NVRL-79IRL02689/2020\|EPI_ISL_73 | ···················· | ···················· | ···················· | ···················· | ···················· | ···················· |
| Y | 4226 | hCoV-19/Ireland/OY-NVRL-76IRL27010/2020\|EPI_ISL_73 | ···················· | ···················· | ···················· | ···················· | ···················· | ···················· |
| Y | 4227 | hCoV-19/England/MILK-C2F931/2020\|EPI_ISL_733801\|20 | ···················· | ···················· | ···················· | ···················· | ···················· | ···················· |
| Y | 4228 | hCoV-19/England/ALDP-C22F0F/2020\|EPI_ISL_733803\|20 | ···················· | ···················· | ···················· | ···················· | ···················· | ···················· |
| Y | 4229 | hCoV-19/England/CAMC-C2D326/2020\|EPI_ISL_733804\|20 | ···················· | ···················· | ···················· | ···················· | ···················· | ···················· |
| Y | 4230 | hCoV-19/England/ALDP-C23029/2020\|EPI_ISL_733806\|20 | ···················· | ···················· | ···················· | ···················· | ···················· | ···················· |
| Y | 4231 | hCoV-19/England/ALDP-C2329C/2020\|EPI_ISL_733807\|20 | ···················· | ···················· | ···················· | ···················· | ···················· | ···················· |
| Y | 4232 | hCoV-19/England/MILK-C2F913/2020\|EPI_ISL_733811\|20 | ···················· | ···················· | ···················· | ···················· | ···················· | ···················· |
| Y | 4233 | hCoV-19/England/MILK-C2FC0B/2020\|EPI_ISL_733825\|20 | ···················· | ···················· | ···················· | ···················· | ···················· | ···················· |
| Y | 4234 | hCoV-19/England/ALDP-C24495/2020\|EPI_ISL_733837\|20 | ···················· | ···················· | ···················· | ···················· | ···················· | ···················· |
| Y | 4235 | hCoV-19/England/MILK-C2FC29/2020\|EPI_ISL_733838\|20 | ···················· | ···················· | ···················· | ···················· | ···················· | ···················· |
| Y | 4236 | hCoV-19/England/ALDP-C22E8A/2020\|EPI_ISL_733844\|20 | ···················· | ···················· | ···················· | ···················· | ···················· | ···················· |
| Y | 4237 | hCoV-19/England/CAMC-C263B4/2020\|EPI_ISL_733846\|20 | ···················· | ···················· | ···················· | ···················· | ···················· | ···················· |
| Y | 4238 | hCoV-19/England/ALDP-C23223/2020\|EPI_ISL_733848\|20 | ···················· | ···················· | ···················· | ···················· | ···················· | ···················· |
| Y | 4239 | hCoV-19/England/CAMC-C264FD/2020\|EPI_ISL_733849\|20 | ···················· | ···················· | ···················· | ···················· | ···················· | ···················· |
| Y | 4240 | hCoV-19/England/MILK-C30584/2020\|EPI_ISL_733851\|20 | ···················· | ···················· | ···················· | ···················· | ·····T·············· | ···················· |
| Y | 4241 | hCoV-19/England/CAMC-C2CE44/2020\|EPI_ISL_733853\|20 | ···················· | ···················· | ···················· | ···················· | ···················· | ···················· |
| Y | 4242 | hCoV-19/England/CAMC-C2CFAB/2020\|EPI_ISL_733854\|20 | ···················· | ···················· | ···················· | ···················· | ···················· | ···················· |
| Y | 4243 | hCoV-19/England/MILK-C2FBA4/2020\|EPI_ISL_733855\|20 | ···················· | ···················· | ···················· | ···················· | ···················· | ···················· |
| Y | 4244 | hCoV-19/England/QEUH-C5D5DC/2020\|EPI_ISL_736003\|20 | ···················· | ···················· | ···················· | ···················· | ···················· | ···················· |
| Y | 4245 | hCoV-19/England/QEUH-C5C731/2020\|EPI_ISL_736007\|20 | ···················· | ···················· | ···················· | ···················· | ···················· | ···················· |
| Y | 4246 | hCoV-19/England/ALDP-C5860B/2020\|EPI_ISL_736008\|20 | ···················· | ···················· | ···················· | ···················· | ···················· | ···················· |
| Y | 4247 | hCoV-19/England/ALDP-C5801C/2020\|EPI_ISL_736010\|20 | ···················· | ···················· | ···················· | ···················· | ···················· | ···················· |
| Y | 4248 | hCoV-19/England/ALDP-C588E7/2020\|EPI_ISL_736011\|20 | ···················· | ···················· | ···················· | ···················· | ···················· | ···················· |
| Y | 4249 | hCoV-19/England/ALDP-C5887E/2020\|EPI_ISL_736012\|20 | ···················· | ···················· | ···················· | ···················· | ···················· | ···················· |
| Y | 4250 | hCoV-19/England/ALDP-C58911/2020\|EPI_ISL_736013\|20 | ···················· | ···················· | ···················· | ···················· | ···················· | ···················· |
| Y | 4251 | hCoV-19/England/ALDP-C59725/2020\|EPI_ISL_736014\|20 | ···················· | ···················· | ···················· | ···················· | ·····T·············· | ···················· |
| Y | 4252 | hCoV-19/England/QEUH-C5CB9F/2020\|EPI_ISL_736015\|20 | ···················· | ···················· | ···················· | ···················· | ···················· | ···················· |
| Y | 4253 | hCoV-19/England/ALDP-C5861A/2020\|EPI_ISL_736016\|20 | ···················· | ···················· | ···················· | ···················· | ···················· | ···················· |
| Y | 4254 | hCoV-19/England/QEUH-C5B802/2020\|EPI_ISL_736018\|20 | ···················· | ···················· | ···················· | ···················· | ···················· | ···················· |
| Y | 4255 | hCoV-19/England/ALDP-C583B9/2020\|EPI_ISL_736020\|20 | ···················· | ···················· | ···················· | ···················· | ·····T·············· | ···················· |
| Y | 4256 | hCoV-19/England/ALDP-C58F3D/2020\|EPI_ISL_736021\|20 | ···················· | ···················· | ···················· | ···················· | ···················· | ···················· |
| Y | 4257 | hCoV-19/England/ALDP-C5889C/2020\|EPI_ISL_736023\|20 | ···················· | ···················· | ···················· | ···················· | ···················· | ···················· |
| Y | 4258 | hCoV-19/England/QEUH-C5D5EB/2020\|EPI_ISL_736024\|20 | ···················· | ···················· | ···················· | ···················· | ···················· | ···················· |
| Y | 4259 | hCoV-19/England/ALDP-C58902/2020\|EPI_ISL_736028\|20 | ···················· | ···················· | ···················· | ···················· | ···················· | ···················· |
| Y | 4260 | hCoV-19/England/ALDP-C585D1/2020\|EPI_ISL_736029\|20 | ···················· | ···················· | ···················· | ···················· | ···················· | ···················· |
| Y | 4261 | hCoV-19/England/ALDP-C58629/2020\|EPI_ISL_736034\|20 | ···················· | ···················· | ···················· | ···················· | ···················· | ···················· |
| Y | 4262 | hCoV-19/England/QEUH-C5BD7C/2020\|EPI_ISL_736038\|20 | ···················· | ···················· | ···················· | ···················· | ···················· | ···················· |
| Y | 4263 | hCoV-19/England/ALDP-C57F3E/2020\|EPI_ISL_736039\|20 | ···················· | ···················· | ···················· | ···················· | ···················· | ···················· |
| Y | 4264 | hCoV-19/England/ALDP-C58A1E/2020\|EPI_ISL_736040\|20 | ···················· | ···················· | ···················· | ···················· | ···················· | ···················· |
| Y | 4265 | hCoV-19/England/QEUH-C5BD8B/2020\|EPI_ISL_736041\|20 | ···················· | ···················· | ···················· | ···················· | ···················· | ···················· |
| Y | 4266 | hCoV-19/England/ALDP-C57F01/2020\|EPI_ISL_736043\|20 | ···················· | ···················· | ···················· | ···················· | ···················· | ···················· |
| Y | 4267 | hCoV-19/England/ALDP-C5843E/2020\|EPI_ISL_736044\|20 | ···················· | ···················· | ···················· | ···················· | ···················· | ···················· |
| Y | 4268 | hCoV-19/England/ALDP-C58A87/2020\|EPI_ISL_736045\|20 | ···················· | ···················· | ···················· | ···················· | ···················· | ···················· |
| Y | 4269 | hCoV-19/England/ALDP-C58BEE/2020\|EPI_ISL_736046\|20 | ···················· | ···················· | ···················· | ···················· | ···················· | ···················· |
| Y | 4270 | hCoV-19/England/ALDP-C57C73/2020\|EPI_ISL_736047\|20 | ···················· | ···················· | ···················· | ···················· | ···················· | ···················· |
| Y | 4271 | hCoV-19/England/ALDP-C58F5B/2020\|EPI_ISL_736048\|20 | ···················· | ···················· | ···················· | ···················· | ···················· | ···················· |
| Y | 4272 | hCoV-19/England/ALDP-C58638/2020\|EPI_ISL_736051\|20 | ···················· | ···················· | ···················· | ···················· | ···················· | ···················· |
| Y | 4273 | hCoV-19/England/ALDP-C5844D/2020\|EPI_ISL_736054\|20 | ···················· | ···················· | ···················· | ···················· | ···················· | ···················· |
| Y | 4274 | hCoV-19/England/ALDP-C580B2/2020\|EPI_ISL_736056\|20 | ···················· | ···················· | ···················· | ···················· | ···················· | ···················· |
| Y | 4275 | hCoV-19/England/QEUH-C5B750/2020\|EPI_ISL_736057\|20 | ···················· | ···················· | ···················· | ···················· | ···················· | ···················· |
| Y | 4276 | hCoV-19/England/ALDP-C58A78/2020\|EPI_ISL_736058\|20 | ···················· | ···················· | ···················· | ···················· | ·····T·············· | ···················· |
| Y | 4277 | hCoV-19/England/QEUH-C5BD12/2020\|EPI_ISL_736059\|20 | ···················· | ···················· | ···················· | ···················· | ···················· | ···················· |
| Y | 4278 | hCoV-19/England/ALDP-C57E5F/2020\|EPI_ISL_736061\|20 | ···················· | ···················· | ···················· | ···················· | ···················· | ···················· |
| Y | 4279 | hCoV-19/England/ALDP-C57C91/2020\|EPI_ISL_736063\|20 | ···················· | ···················· | ···················· | ···················· | ···················· | ···················· |
| Y | 4280 | hCoV-19/England/ALDP-C585E0/2020\|EPI_ISL_736065\|20 | ···················· | ···················· | ···················· | ···················· | ···················· | ···················· |
| Y | 4281 | hCoV-19/England/QEUH-C5B76F/2020\|EPI_ISL_736066\|20 | ···················· | ···················· | ···················· | ···················· | ···················· | ···················· |
| Y | 4282 | hCoV-19/England/ALDP-C58137/2020\|EPI_ISL_736070\|20 | ···················· | ·A·················· | ···················· | ···················· | ···················· | ··A················· |
| Y | 4283 | hCoV-19/England/ALDP-C581DD/2020\|EPI_ISL_736071\|20 | ···················· | ···················· | ···················· | ···················· | ···················· | ···················· |
| Y | 4284 | hCoV-19/England/QEUH-C5D509/2020\|EPI_ISL_736072\|20 | ···················· | ···················· | ···················· | ···················· | ···················· | ···················· |
| Y | 4285 | hCoV-19/England/ALDP-C586DE/2020\|EPI_ISL_736073\|20 | ···················· | ···A················ | ···················· | ···················· | ···················· | ···················· |

Y 4286 hCoV-19/England/ALDP-C57D9E/2020|EPI_ISL_736074|20 ···················· ···················· ···················· ···················· ···················· ····················
Y 4287 hCoV-19/England/ALDP-C5987D/2020|EPI_ISL_736076|20 ···················· ···················· ···················· ···················· ·····T·············· ····················
Y 4288 hCoV-19/England/ALDP-C5846B/2020|EPI_ISL_736077|20 ···················· ···················· ···················· ···················· ···················· ····················
Y 4289 hCoV-19/England/ALDP-C57CEC/2020|EPI_ISL_736079|20 ···················· ···················· ···················· ···················· ···················· ····················
Y 4290 hCoV-19/England/ALDP-C57CA0/2020|EPI_ISL_736080|20 ···················· ···················· ···················· ···················· ···················· ····················
Y 4291 hCoV-19/England/ALDP-C59163/2020|EPI_ISL_736081|20 ···················· ···················· ···················· ···················· ···················· ····················
Y 4292 hCoV-19/England/ALDP-C58595/2020|EPI_ISL_736083|20 ···················· ···················· ···················· ···················· ···················· ····················
Y 4293 hCoV-19/England/ALDP-C58216/2020|EPI_ISL_736084|20 ···················· ···················· ···················· ···················· ···················· ····················
Y 4294 hCoV-19/England/ALDP-C57C64/2020|EPI_ISL_736087|20 ···················· ···················· ···················· ···················· ···················· ····················
Y 4295 hCoV-19/England/ALDP-C59743/2020|EPI_ISL_736089|20 ···················· ···················· ···················· ···················· ···················· ····················
Y 4297 hCoV-19/USA/MI-MDHHS-SC22688/2020|EPI_ISL_737302|2 ···················· ···················· ···················· ···················· ···················· ····················
Y 4298 hCoV-19/USA/MI-MDHHS-SC22689/2020|EPI_ISL_737303|2 ···················· ···················· ···················· ···················· ···················· ····················
Y 4299 hCoV-19/USA/MI-MDHHS-SC22690/2020|EPI_ISL_737304|2 ···················· ···················· ···················· ···················· ···················· ····················
Y 4300 hCoV-19/USA/MI-MDHHS-SC22691/2020|EPI_ISL_737305|2 ···················· ·A·················· ···················· ···················· ···················· ····················
Y 4301 hCoV-19/Belgium/ULG-11058/2020|EPI_ISL_737310|2020 ···················· ···················· ···················· ···················· ···················· ····················
Y 4302 hCoV-19/Belgium/ULG-11059/2020|EPI_ISL_737311|2020 ···················· ···················· ···················· ···················· ···················· ····················
Y 4303 hCoV-19/Belgium/ULG-11062/2020|EPI_ISL_737313|2020 ···················· ···A················ ···················· ···················· ···················· ····················
Y 4304 hCoV-19/Belgium/ULG-11063/2020|EPI_ISL_737314|2020 ···················· ···················· ···················· ···················· ···················· ····················
Y 4305 hCoV-19/Belgium/ULG-11068/2020|EPI_ISL_737319|2020 ···················· ···················· ···················· ···················· ···················· ····················
Y 4306 hCoV-19/Belgium/ULG-11069/2020|EPI_ISL_737320|2020 ···················· ···················· ···················· ···················· ·····T·············· ····················
Y 4307 hCoV-19/Belgium/ULG-11071/2020|EPI_ISL_737322|2020 ···················· ···················· ···················· ···················· ···················· ····················
Y 4308 hCoV-19/Belgium/ULG-11074/2020|EPI_ISL_737325|2020 ···················· ···················· ···················· ···················· ···················· ····················
Y 4309 hCoV-19/Belgium/ULG-11075/2020|EPI_ISL_737326|2020 ···················· ···················· ···················· ···················· ···················· ····················
Y 4310 hCoV-19/Belgium/ULG-11079/2020|EPI_ISL_737329|2020 ···················· ···A················ ···················· ···················· ···················· ····················
Y 4311 hCoV-19/Belgium/ULG-11080/2020|EPI_ISL_737330|2020 ···················· ···················· ···················· ···················· ···················· ····················
Y 4312 hCoV-19/Belgium/ULG-11083/2020|EPI_ISL_737333|2020 ···················· ···A················ ···················· ···················· ···················· ····················
Y 4313 hCoV-19/Belgium/ULG-11085/2020|EPI_ISL_737335|2020 ···················· ···················· ···················· ···················· ···················· ····················
Y 4314 hCoV-19/Belgium/ULG-11089/2020|EPI_ISL_737338|2020 ···················· ···A················ ···················· ···················· ···················· ····················
Y 4315 hCoV-19/Belgium/ULG-11094/2020|EPI_ISL_737343|2020 ···················· ···················· ···················· ···················· ···················· ····················
Y 4316 hCoV-19/Belgium/ULG-11095/2020|EPI_ISL_737344|2020 ···················· ···················· ···················· ···················· ···················· ····················
Y 4317 hCoV-19/Belgium/ULG-11097/2020|EPI_ISL_737346|2020 ···················· ···················· ···················· ···················· ···················· ····················
Y 4318 hCoV-19/Belgium/ULG-11099/2020|EPI_ISL_737347|2020 ···················· ···A················ ···················· ···················· ···················· ····················
Y 4319 hCoV-19/Belgium/ULG-11101/2020|EPI_ISL_737349|2020 ···················· ···················· ···················· ···················· ···················· ····················
Y 4320 hCoV-19/Belgium/ULG-11102/2020|EPI_ISL_737350|2020 ···················· ···················· ···················· ···················· ···················· ····················
Y 4321 hCoV-19/Belgium/ULG-11103/2020|EPI_ISL_737351|2020 ···················· ···················· ···················· ···················· ···················· ····················
Y 4322 hCoV-19/Belgium/ULG-11105/2020|EPI_ISL_737353|2020 ···················· ···················· ···················· ···················· ···················· ····················
Y 4323 hCoV-19/Belgium/ULG-11106/2020|EPI_ISL_737354|2020 ···················· ···················· ···················· ···················· ···················· ····················
Y 4324 hCoV-19/Belgium/ULG-11109/2020|EPI_ISL_737357|2020 ···················· ···················· ···················· ···················· ·····T·············· ····················
Y 4325 hCoV-19/Belgium/ULG-11111/2020|EPI_ISL_737359|2020 ···················· ···················· ···················· ···················· ·····T·············· ····················
Y 4326 hCoV-19/Belgium/ULG-11112/2020|EPI_ISL_737360|2020 ···················· ···················· ···················· ···················· ·····T·············· ····················
Y 4327 hCoV-19/Belgium/ULG-11114/2020|EPI_ISL_737362|2020 ···················· ···················· ···················· ···················· ···················· ····················
Y 4328 hCoV-19/Belgium/ULG-11115/2020|EPI_ISL_737363|2020 ···················· ···················· ···················· ···················· ···················· ····················
Y 4329 hCoV-19/Belgium/ULG-11116/2020|EPI_ISL_737364|2020 ···················· ···················· ···················· ···················· ···················· ····················
Y 4330 hCoV-19/Belgium/ULG-11117/2020|EPI_ISL_737365|2020 ···················· ···················· ···················· ···················· ···················· ····················
Y 4331 hCoV-19/Belgium/ULG-11119/2020|EPI_ISL_737366|2020 ···················· ···················· ···················· ···················· ···················· ····················
Y 4332 hCoV-19/Belgium/ULG-11120/2020|EPI_ISL_737367|2020 ···················· ···················· ···················· ···················· ·····T·············· ····················
Y 4333 hCoV-19/Belgium/ULG-11121/2020|EPI_ISL_737368|2020 ···················· ···················· ···················· ···················· ···················· ····················
Y 4334 hCoV-19/Belgium/ULG-11122/2020|EPI_ISL_737369|2020 ···················· ···················· ···················· ···················· ···················· ····················
Y 4335 hCoV-19/Belgium/ULG-11123/2020|EPI_ISL_737370|2020 ···················· ···················· ···················· ···················· ···················· ····················
Y 4336 hCoV-19/Belgium/ULG-11124/2020|EPI_ISL_737371|2020 ···················· ···················· ···················· ···················· ···················· ····················
Y 4337 hCoV-19/Belgium/ULG-11128/2020|EPI_ISL_737372|2020 ···················· ···················· ···················· ···················· ···················· ····················
Y 4338 hCoV-19/Belgium/ULG-11129/2020|EPI_ISL_737373|2020 ···················· ···················· ···················· ···················· ·····T·············· ····················
Y 4339 hCoV-19/Belgium/ULG-11132/2020|EPI_ISL_737376|2020 ···················· ···················· ···················· ···················· ·····T·············· ····················
Y 4340 hCoV-19/Belgium/ULG-11133/2020|EPI_ISL_737377|2020 ···················· ···················· ···················· ···················· ···················· ····················
Y 4341 hCoV-19/Belgium/ULG-11135/2020|EPI_ISL_737379|2020 ···················· ···················· ···················· ···················· ···················· ····················
Y 4342 hCoV-19/Belgium/ULG-11137/2020|EPI_ISL_737381|2020 ···················· ···················· ···················· ···················· ···················· ····················
Y 4343 hCoV-19/Belgium/ULG-11139/2020|EPI_ISL_737383|2020 ···················· ···················· ···················· ···················· ···················· ····················
Y 4344 hCoV-19/Belgium/ULG-11140/2020|EPI_ISL_737384|2020 ···················· ···················· ···················· ···················· ···················· ····················
Y 4345 hCoV-19/Belgium/ULG-11141/2020|EPI_ISL_737385|2020 ···················· ···················· ···················· ···················· ···················· ····················
Y 4346 hCoV-19/Belgium/ULG-11142/2020|EPI_ISL_737386|2020 ···················· ···················· ···················· ···················· ···················· ····················
Y 4347 hCoV-19/Belgium/ULG-11143/2020|EPI_ISL_737387|2020 ···················· ···················· ···················· ···················· ···················· ····················
Y 4348 hCoV-19/Belgium/ULG-11146/2020|EPI_ISL_737390|2020 ···················· ···················· ···················· ···················· ···················· ····················
Y 4349 hCoV-19/Belgium/ULG-11147/2020|EPI_ISL_737391|2020 ···················· ···················· ···················· ···················· ···················· ····················
Y 4350 hCoV-19/Belgium/ULG-11148/2020|EPI_ISL_737392|2020 ···················· ···················· ···················· ···················· ···················· ····················
Y 4351 hCoV-19/Belgium/ULG-11149/2020|EPI_ISL_737393|2020 ···················· ···················· ···················· ···················· ···················· ····················
Y 4352 hCoV-19/Belgium/ULG-11150/2020|EPI_ISL_737394|2020 ···················· ···················· ···················· ···················· ···················· ····················
Y 4353 hCoV-19/England/NORW-F987B/2020|EPI_ISL_725400|202 ···················· ···················· ···················· ···················· ···················· ····················
Y 4354 hCoV-19/England/NORW-F988A/2020|EPI_ISL_725401|202 ···················· ···················· ···················· ···················· ···················· ····················
Y 4355 hCoV-19/England/NORW-F98A8/2020|EPI_ISL_725403|202 ···················· ···················· ···················· ···················· ···················· ····················
Y 4356 hCoV-19/England/NORW-F98C6/2020|EPI_ISL_725405|202 ···················· ···················· ···················· ···················· ···················· ····················
Y 4357 hCoV-19/England/NORW-F98D5/2020|EPI_ISL_725406|202 ···················· ·A·················· ···················· ···················· ···················· ··A·················
Y 4358 hCoV-19/England/NORW-F98F3/2020|EPI_ISL_725408|202 ···················· ···················· ···················· ···················· ···················· ····················
Y 4359 hCoV-19/England/NORW-F991E/2020|EPI_ISL_725409|202 ···················· ···················· ···················· ···················· ···················· ····················
Y 4362 hCoV-19/England/NORW-F9969/2020|EPI_ISL_725413|202 ···················· ···················· ···················· ···················· ···················· ····················
Y 4363 hCoV-19/England/NORW-F9978/2020|EPI_ISL_725414|202 ···················· ···················· ···················· ···················· ···················· ····················
Y 4364 hCoV-19/England/NORW-F99C3/2020|EPI_ISL_725415|202 ···················· ···················· ···················· ···················· ···················· ····················
Y 4365 hCoV-19/England/NORW-F99E1/2020|EPI_ISL_725416|202 ···················· ···················· ···················· ···················· ···················· ····················
Y 4366 hCoV-19/England/NORW-F9ADF/2020|EPI_ISL_725424|202 ···················· ···················· ···················· ···················· ···················· ····················
Y 4367 hCoV-19/England/NORW-F9B36/2020|EPI_ISL_725428|202 ···················· ···················· ···················· ···················· ···················· ····················
Y 4368 hCoV-19/England/NORW-F9B54/2020|EPI_ISL_725429|202 ···················· ···················· ···················· ···················· ···················· ····················

| | | | | | | | | |
|---|---|---|---|---|---|---|---|---|
| Y | 4369 | hCoV-19/England/NORW-F9B63/2020\|EPI_ISL_725430\|202 | .................... | .................... | .................... | .................... | .................... | .................... |
| Y | 4370 | hCoV-19/England/NORW-F9B72/2020\|EPI_ISL_725431\|202 | .................... | .................... | .................... | .................... | .................... | .................... |
| Y | 4371 | hCoV-19/England/NORW-F9C15/2020\|EPI_ISL_725434\|202 | .................... | .................... | .................... | .................... | .................... | .................... |
| Y | 4372 | hCoV-19/England/NORW-F9C42/2020\|EPI_ISL_725435\|202 | .................... | .................... | .................... | .................... | .................... | .................... |
| Y | 4373 | hCoV-19/England/NORW-F9C51/2020\|EPI_ISL_725436\|202 | .................... | .................... | .................... | .................... | .................... | .................... |
| Y | 4374 | hCoV-19/England/NORW-F9C60/2020\|EPI_ISL_725437\|202 | .................... | .................... | .................... | .................... | .................... | .................... |
| Y | 4375 | hCoV-19/England/NORW-F9CAC/2020\|EPI_ISL_725439\|202 | .................... | .................... | .................... | .................... | .................... | .................... |
| Y | 4376 | hCoV-19/England/NORW-FA027/2020\|EPI_ISL_725460\|202 | .................... | .................... | ..A................. | .................... | .................... | .................... |
| Y | 4377 | hCoV-19/England/NORW-FA063/2020\|EPI_ISL_725463\|202 | .................... | .................... | .................... | .................... | .................... | .................... |
| Y | 4378 | hCoV-19/England/NORW-FA081/2020\|EPI_ISL_725464\|202 | .................... | .................... | .................... | .................... | .................... | .................... |
| Y | 4379 | hCoV-19/England/NORW-FA0BE/2020\|EPI_ISL_725466\|202 | .................... | .................... | .................... | .................... | .................... | .................... |
| Y | 4380 | hCoV-19/England/NORW-FA0CD/2020\|EPI_ISL_725467\|202 | .................... | .A.................. | .................... | .................... | .................... | ..A................. |
| Y | 4383 | hCoV-19/Wales/PHWC-496C16/2020\|EPI_ISL_726703\|2020 | .................... | .................... | .................... | .................... | .................... | .................... |
| Y | 4385 | hCoV-19/Wales/PHWC-496C34/2020\|EPI_ISL_726705\|2020 | .................... | .................... | .................... | .................... | .................... | .................... |
| Y | 4386 | hCoV-19/Wales/PHWC-496C43/2020\|EPI_ISL_726706\|2020 | .................... | .................... | .................... | .................... | .................... | .................... |
| Y | 4388 | hCoV-19/Wales/PHWC-496C61/2020\|EPI_ISL_726708\|2020 | .................... | .................... | .................... | .................... | .................... | .................... |
| Y | 4389 | hCoV-19/Wales/PHWC-496C8F/2020\|EPI_ISL_726709\|2020 | .................... | .................... | .................... | .................... | .................... | .................... |
| Y | 4391 | hCoV-19/Wales/PHWC-496CBC/2020\|EPI_ISL_726712\|2020 | .................... | .................... | .................... | .................... | .................... | .................... |
| Y | 4393 | hCoV-19/Wales/PHWC-496CDA/2020\|EPI_ISL_726714\|2020 | .................... | .................... | .................... | .................... | .................... | .................... |
| Y | 4394 | hCoV-19/Wales/PHWC-496CE9/2020\|EPI_ISL_726715\|2020 | .................... | .................... | .................... | .................... | .................... | .................... |
| Y | 4395 | hCoV-19/Wales/PHWC-496CF8/2020\|EPI_ISL_726716\|2020 | .................... | .................... | .................... | .................... | .................... | .................... |
| Y | 4396 | hCoV-19/Wales/PHWC-496D13/2020\|EPI_ISL_726718\|2020 | .................... | .................... | .................... | .................... | .................... | .................... |
| Y | 4398 | hCoV-19/Wales/PHWC-496D31/2020\|EPI_ISL_726720\|2020 | .................... | .................... | .................... | .................... | .................... | .................... |
| Y | 4399 | hCoV-19/Wales/PHWC-496D40/2020\|EPI_ISL_726721\|2020 | .................... | .................... | .................... | .................... | .................... | .................... |
| Y | 4400 | hCoV-19/Wales/PHWC-496D5F/2020\|EPI_ISL_726722\|2020 | .................... | .................... | .................... | .................... | .................... | .................... |
| Y | 4401 | hCoV-19/Wales/PHWC-496D6E/2020\|EPI_ISL_726723\|2020 | .................... | .................... | .................... | .................... | .................... | .................... |
| Y | 4402 | hCoV-19/Wales/PHWC-496D7D/2020\|EPI_ISL_726724\|2020 | .................... | .................... | .................... | .................... | .................... | .................... |
| Y | 4403 | hCoV-19/Wales/PHWC-496D8C/2020\|EPI_ISL_726725\|2020 | .................... | .................... | .................... | .................... | .................... | .................... |
| Y | 4404 | hCoV-19/Wales/PHWC-496D9B/2020\|EPI_ISL_726726\|2020 | .................... | .................... | .................... | .................... | .................... | .................... |
| Y | 4406 | hCoV-19/Wales/PHWC-496DB9/2020\|EPI_ISL_726728\|2020 | .................... | .................... | .................... | .................... | .................... | .................... |
| Y | 4407 | hCoV-19/Wales/PHWC-496DC8/2020\|EPI_ISL_726729\|2020 | .................... | .................... | .................... | .................... | .................... | .................... |
| Y | 4408 | hCoV-19/Wales/PHWC-496DE6/2020\|EPI_ISL_726731\|2020 | .................... | .................... | .................... | .................... | .................... | .................... |
| Y | 4410 | hCoV-19/Wales/PHWC-496E01/2020\|EPI_ISL_726733\|2020 | .................... | ......A............. | .................... | .................... | .................... | .................... |
| Y | 4411 | hCoV-19/Wales/PHWC-496E10/2020\|EPI_ISL_726734\|2020 | .................... | ......A............. | .................... | .................... | .................... | .................... |
| Y | 4412 | hCoV-19/Wales/PHWC-496E3E/2020\|EPI_ISL_726735\|2020 | .................... | .................... | .................... | .................... | .................... | .................... |
| Y | 4413 | hCoV-19/Wales/PHWC-496E4D/2020\|EPI_ISL_726736\|2020 | .................... | .................... | .................... | .................... | .................... | .................... |
| Y | 4414 | hCoV-19/Wales/PHWC-496E5C/2020\|EPI_ISL_726737\|2020 | .................... | .................... | .................... | .................... | .................... | .................... |
| Y | 4417 | hCoV-19/Wales/PHWC-496E89/2020\|EPI_ISL_726740\|2020 | .................... | .................... | .................... | .................... | .................... | .................... |
| Y | 4418 | hCoV-19/Wales/PHWC-496EA7/2020\|EPI_ISL_726742\|2020 | .................... | .................... | .................... | .................... | .................... | .................... |
| Y | 4419 | hCoV-19/Wales/PHWC-496EB6/2020\|EPI_ISL_726743\|2020 | .................... | .................... | .................... | .................... | .................... | .................... |
| Y | 4423 | hCoV-19/Wales/PHWC-496F0E/2020\|EPI_ISL_726748\|2020 | .................... | .................... | .................... | .................... | .................... | .................... |
| Y | 4424 | hCoV-19/Wales/PHWC-496F1D/2020\|EPI_ISL_726749\|2020 | .................... | .................... | .................... | .................... | .................... | .................... |
| Y | 4426 | hCoV-19/Wales/PHWC-496F3B/2020\|EPI_ISL_726751\|2020 | .................... | .................... | .................... | .................... | .................... | .................... |
| Y | 4427 | hCoV-19/Wales/PHWC-496F59/2020\|EPI_ISL_726753\|2020 | .................... | .................... | .................... | .................... | .................... | .................... |
| Y | 4428 | hCoV-19/Wales/PHWC-496F68/2020\|EPI_ISL_726754\|2020 | .................... | .................... | .................... | .................... | .................... | .................... |
| Y | 4429 | hCoV-19/Wales/PHWC-496F77/2020\|EPI_ISL_726755\|2020 | .................... | .................... | .................... | .................... | .................... | .................... |
| Y | 4431 | hCoV-19/Wales/PHWC-496F95/2020\|EPI_ISL_726757\|2020 | .................... | .................... | .................... | .................... | .................... | .................... |
| Y | 4432 | hCoV-19/Wales/PHWC-496FB3/2020\|EPI_ISL_726759\|2020 | .................... | .................... | .................... | .................... | .................... | .................... |
| Y | 4433 | hCoV-19/Wales/PHWC-496FC2/2020\|EPI_ISL_726760\|2020 | .................... | .................... | .................... | .................... | .................... | .................... |
| Y | 4434 | hCoV-19/Wales/PHWC-496FD1/2020\|EPI_ISL_726761\|2020 | .................... | .................... | .................... | .................... | .................... | .................... |
| Y | 4438 | hCoV-19/Wales/PHWC-497019/2020\|EPI_ISL_726765\|2020 | .................... | .................... | .................... | .................... | .................... | .................... |
| Y | 4441 | hCoV-19/Wales/PHWC-497046/2020\|EPI_ISL_726768\|2020 | .................... | .................... | .................... | .................... | .................... | .................... |
| Y | 4442 | hCoV-19/Wales/PHWC-497055/2020\|EPI_ISL_726769\|2020 | .................... | .................... | .................... | .................... | .................... | .................... |
| Y | 4443 | hCoV-19/Wales/PHWC-4971AD/2020\|EPI_ISL_726790\|2020 | .................... | .................... | .................... | .................... | .................... | .................... |
| Y | 4444 | hCoV-19/Wales/PHWC-4971BC/2020\|EPI_ISL_726791\|2020 | .................... | .................... | .................... | .................... | .................... | .................... |
| Y | 4445 | hCoV-19/Wales/PHWC-4971CB/2020\|EPI_ISL_726792\|2020 | .................... | .................... | .................... | .................... | .................... | .................... |
| Y | 4446 | hCoV-19/Wales/PHWC-4971DA/2020\|EPI_ISL_726793\|2020 | .................... | .................... | .................... | .................... | .................... | .................... |
| Y | 4447 | hCoV-19/Wales/PHWC-4971E9/2020\|EPI_ISL_726794\|2020 | .................... | .................... | .................... | .................... | .................... | .................... |
| Y | 4448 | hCoV-19/Wales/PHWC-4971F8/2020\|EPI_ISL_726795\|2020 | .................... | .................... | .................... | .................... | .................... | .................... |
| Y | 4449 | hCoV-19/Wales/PHWC-497204/2020\|EPI_ISL_726796\|2020 | .................... | .................... | .................... | .................... | .................... | .................... |
| Y | 4453 | hCoV-19/England/ALDP-C39BD4/2020\|EPI_ISL_731200\|20 | .................... | .................... | .................... | .................... | .................... | .................... |
| Y | 4454 | hCoV-19/England/ALDP-C3C98C/2020\|EPI_ISL_731201\|20 | .................... | .................... | .................... | .................... | .................... | .................... |
| Y | 4455 | hCoV-19/England/ALDP-C3C430/2020\|EPI_ISL_731202\|20 | .................... | .................... | .................... | .................... | .................... | .................... |
| Y | 4456 | hCoV-19/England/ALDP-C3C8BC/2020\|EPI_ISL_731203\|20 | .................... | .................... | .................... | .................... | .................... | .................... |
| Y | 4457 | hCoV-19/England/ALDP-C3A371/2020\|EPI_ISL_731204\|20 | .................... | .................... | .................... | .................... | .................... | .................... |
| Y | 4458 | hCoV-19/England/ALDP-C3C4C7/2020\|EPI_ISL_731205\|20 | .................... | .................... | .................... | .................... | .................... | .................... |
| Y | 4459 | hCoV-19/England/ALDP-C3D086/2020\|EPI_ISL_731207\|20 | .................... | .................... | .................... | .................... | .................... | .................... |
| Y | 4460 | hCoV-19/England/ALDP-C3B765/2020\|EPI_ISL_731208\|20 | .................... | .................... | .................... | .................... | .................... | .................... |
| Y | 4461 | hCoV-19/England/ALDP-C3B817/2020\|EPI_ISL_731209\|20 | .................... | .................... | .................... | .................... | .................... | .................... |
| Y | 4462 | hCoV-19/England/ALDP-C3C4D6/2020\|EPI_ISL_731211\|20 | .................... | .................... | .................... | .................... | .................... | .................... |
| Y | 4463 | hCoV-19/England/ALDP-C3BBC3/2020\|EPI_ISL_731212\|20 | .................... | .................... | .................... | .................... | .................... | .................... |
| Y | 4464 | hCoV-19/England/ALDP-C3DB58/2020\|EPI_ISL_731213\|20 | .................... | .................... | .................... | .................... | .................... | .................... |
| Y | 4465 | hCoV-19/England/ALDP-C3BDAF/2020\|EPI_ISL_731216\|20 | .................... | .................... | .................... | .................... | .................... | .................... |
| Y | 4466 | hCoV-19/England/ALDP-C3C245/2020\|EPI_ISL_731219\|20 | .................... | .................... | .................... | .................... | .................... | .................... |
| Y | 4467 | hCoV-19/England/ALDP-C3CC47/2020\|EPI_ISL_731220\|20 | .................... | .................... | .................... | .................... | .................... | .................... |
| Y | 4468 | hCoV-19/England/ALDP-C3D98B/2020\|EPI_ISL_731221\|20 | .................... | .................... | .................... | .................... | .................... | .................... |
| Y | 4469 | hCoV-19/England/ALDP-C3B7ED/2020\|EPI_ISL_731224\|20 | .................... | .................... | .................... | .................... | .................... | .................... |
| Y | 4470 | hCoV-19/England/ALDP-C3BC1B/2020\|EPI_ISL_731229\|20 | .................... | .................... | .................... | .................... | .................... | .................... |
| Y | 4471 | hCoV-19/England/ALDP-C3D921/2020\|EPI_ISL_731230\|20 | .................... | .................... | .................... | .................... | .................... | .................... |
| Y | 4472 | hCoV-19/England/ALDP-C39C86/2020\|EPI_ISL_731233\|20 | .................... | .................... | .................... | .................... | .................... | .................... |

| | | | | | | | |
|---|---|---|---|---|---|---|---|
| Y | 4473 hCoV-19/England/ALDP-C3C62B/2020\|EPI_ISL_731234\|20 | ···················· | ···················· | ····················· | ···················· | ···················· | ··A················· |
| Y | 4474 hCoV-19/England/ALDP-C3D48A/2020\|EPI_ISL_731236\|20 | ···················· | ···················· | ····················· | ···················· | ···················· | ···················· |
| Y | 4475 hCoV-19/England/ALDP-C3C922/2020\|EPI_ISL_731237\|20 | ···················· | ···················· | ····················· | ···················· | ···················· | ···················· |
| Y | 4476 hCoV-19/England/ALDP-C39CB3/2020\|EPI_ISL_731240\|20 | ···················· | ···················· | ····················· | ···················· | ···················· | ···················· |
| Y | 4477 hCoV-19/England/ALDP-C3C782/2020\|EPI_ISL_731241\|20 | ···················· | ···················· | ····················· | ···················· | ···················· | ···················· |
| Y | 4478 hCoV-19/England/ALDP-C3C1DF/2020\|EPI_ISL_731242\|20 | ···················· | ···················· | ····················· | ···················· | ···················· | ···················· |
| Y | 4479 hCoV-19/England/ALDP-C3C579/2020\|EPI_ISL_731243\|20 | ···················· | ···················· | ····················· | ···················· | ···················· | ···············A···· |
| Y | 4480 hCoV-19/England/ALDP-C3CED8/2020\|EPI_ISL_731245\|20 | ···················· | ···················· | ····················· | ···················· | ···················· | ···················· |
| Y | 4481 hCoV-19/England/ALDP-C3CC38/2020\|EPI_ISL_731246\|20 | ···················· | ······R············· | ····················· | ···················· | ···················· | ···················· |
| Y | 4482 hCoV-19/England/ALDP-C3A229/2020\|EPI_ISL_731248\|20 | ···················· | ···················· | ····················· | ···················· | ···················· | ···················· |
| Y | 4483 hCoV-19/England/ALDP-C3B853/2020\|EPI_ISL_731249\|20 | ···················· | ···················· | ····················· | ···················· | ···················· | ···················· |
| Y | 4484 hCoV-19/England/MILK-BB1021/2020\|EPI_ISL_731251\|20 | ···················· | ···················· | ····················· | ···················· | ···················· | ···················· |
| Y | 4485 hCoV-19/England/ALDP-C3A1E0/2020\|EPI_ISL_731254\|20 | ···················· | ···················· | ····················· | ···················· | ···················· | ···················· |
| Y | 4486 hCoV-19/England/QEUH-C30EA7/2020\|EPI_ISL_731256\|20 | ···················· | ···················· | ····················· | ···················· | ···················· | ···················· |
| Y | 4487 hCoV-19/England/QEUH-C35FCD/2020\|EPI_ISL_731257\|20 | ···················· | ···················· | ····················· | ···················· | ···················· | ···················· |
| Y | 4488 hCoV-19/England/QEUH-C31EC4/2020\|EPI_ISL_731259\|20 | ···················· | ···················· | ····················· | ···················· | ···················· | ···················· |
| Y | 4489 hCoV-19/England/QEUH-C315ED/2020\|EPI_ISL_731261\|20 | ···················· | ···················· | ····················· | ···················· | ···················· | ··········T········· |
| Y | 4490 hCoV-19/England/QEUH-C30A94/2020\|EPI_ISL_731262\|20 | ···················· | ·A·················· | ····················· | ···················· | ···················· | ··A················· |
| Y | 4491 hCoV-19/England/QEUH-C3125F/2020\|EPI_ISL_731264\|20 | ···················· | ···················· | ····················· | ···················· | ···················· | ···················· |
| Y | 4492 hCoV-19/England/ALDP-C3AD91/2020\|EPI_ISL_731265\|20 | ···················· | ···················· | ····················· | ···················· | ···················· | ···················· |
| Y | 4493 hCoV-19/England/QEUH-C3137A/2020\|EPI_ISL_731267\|20 | ···················· | ···················· | ····················· | ···················· | ···················· | ···················· |
| Y | 4494 hCoV-19/England/ALDP-C3A6F0/2020\|EPI_ISL_731268\|20 | ···················· | ···················· | ····················· | ···················· | ···················· | ···················· |
| Y | 4495 hCoV-19/England/ALDP-C3A739/2020\|EPI_ISL_731269\|20 | ···················· | ···················· | ····················· | ···················· | ···················· | ···················· |
| Y | 4496 hCoV-19/England/QEUH-C3134D/2020\|EPI_ISL_731273\|20 | ···················· | ···················· | ····················· | ···················· | ···················· | ···················· |
| Y | 4497 hCoV-19/England/ALDP-C3A4D8/2020\|EPI_ISL_731274\|20 | ···················· | ···················· | ····················· | ···················· | ···················· | ···················· |
| Y | 4498 hCoV-19/England/ALDP-C3AD55/2020\|EPI_ISL_731278\|20 | ···················· | ···················· | ····················· | ···················· | ···················· | ···················· |
| Y | 4499 hCoV-19/England/ALDP-C3A775/2020\|EPI_ISL_731279\|20 | ···················· | ···················· | ····················· | ···················· | ···················· | ···················· |
| Y | 4500 hCoV-19/England/QEUH-C3177E/2020\|EPI_ISL_731280\|20 | ···················· | ···················· | ····················· | ···················· | ···················· | ···················· |
| Y | 4501 hCoV-19/England/QEUH-C30EF2/2020\|EPI_ISL_731281\|20 | ···················· | ···················· | ····················· | ···················· | ···················· | ···················· |
| Y | 4502 hCoV-19/England/QEUH-C313D4/2020\|EPI_ISL_731284\|20 | ···················· | ···················· | ····················· | ···················· | ···················· | ···················· |
| Y | 4503 hCoV-19/England/ALDP-C3922A/2020\|EPI_ISL_731285\|20 | ···················· | ···················· | ····················· | ···················· | ···················· | ···················· |
| Y | 4504 hCoV-19/England/QEUH-C3169F/2020\|EPI_ISL_731286\|20 | ···················· | ···················· | ····················· | ···················· | ···················· | ···················· |
| Y | 4505 hCoV-19/England/QEUH-C315DE/2020\|EPI_ISL_731287\|20 | ···················· | ···················· | ····················· | ···················· | ···················· | ···················· |
| Y | 4506 hCoV-19/England/ALDP-C3ACD0/2020\|EPI_ISL_731288\|20 | ···················· | ···················· | ····················· | ···················· | ···················· | ···················· |
| Y | 4507 hCoV-19/England/ALDP-C39248/2020\|EPI_ISL_731289\|20 | ···················· | ···················· | ····················· | ···················· | ···················· | ···················· |
| Y | 4508 hCoV-19/England/QEUH-C35F36/2020\|EPI_ISL_731291\|20 | ···················· | ···················· | ····················· | ···················· | ······T············· | ···················· |
| Y | 4509 hCoV-19/England/ALDP-C39363/2020\|EPI_ISL_731292\|20 | ···················· | ···················· | ····················· | ···················· | ···················· | ···················· |
| Y | 4510 hCoV-19/England/QEUH-C31662/2020\|EPI_ISL_731295\|20 | ···················· | ···················· | ····················· | ···················· | ···················· | ···················· |
| Y | 4511 hCoV-19/England/ALDP-C39169/2020\|EPI_ISL_731298\|20 | ···················· | ···················· | ····················· | ···················· | ···················· | ···················· |
| Y | 4512 hCoV-19/England/QEUH-C316AE/2020\|EPI_ISL_731299\|20 | ···················· | ···················· | ····················· | ···················· | ···················· | ···················· |
| Y | 4513 hCoV-19/Ireland/D-NVRL-78IRL92236/2020\|EPI_ISL_732 | ···················· | ···················· | ····················· | ···················· | ···················· | ···················· |
| Y | 4514 hCoV-19/Ireland/D-NVRL-78IRL92241/2020\|EPI_ISL_732 | ···················· | ···················· | ····················· | ···················· | ···················· | ···················· |
| Y | 4515 hCoV-19/Ireland/LH-NVRL-20ILR11475/2020\|EPI_ISL_73 | ···················· | ···················· | ····················· | ···················· | ···················· | ···················· |
| Y | 4516 hCoV-19/Germany/BY-IMB06496/2020\|EPI_ISL_732533\|20 | ···················· | ···················· | ····················· | ···················· | ···················· | ···················· |
| Y | 4517 hCoV-19/Germany/BY-IMB08291/2020\|EPI_ISL_732534\|20 | ···················· | ···················· | ····················· | ···················· | ···················· | ···················· |
| Y | 4518 hCoV-19/Germany/BY-IMB07884/2020\|EPI_ISL_732535\|20 | ···················· | ···················· | ····················· | ···················· | ···················· | ···················· |
| Y | 4519 hCoV-19/Germany/BY-IMB07990/2020\|EPI_ISL_732536\|20 | ···················· | ···················· | ····················· | ···················· | ···················· | ···················· |
| Y | 4520 hCoV-19/Germany/BY-IMB07597/2020\|EPI_ISL_732537\|20 | ···················· | ···················· | ····················· | ···················· | ···················· | ···················· |
| Y | 4521 hCoV-19/Germany/BY-IMB06600/2020\|EPI_ISL_732538\|20 | ···················· | ···················· | ····················· | ···················· | ···················· | ···················· |
| Y | 4522 hCoV-19/Germany/BY-IMB06861/2020\|EPI_ISL_732539\|20 | ···················· | ···················· | ····················· | ···················· | ···················· | ···················· |
| Y | 4523 hCoV-19/Germany/BY-IMB07324/2020\|EPI_ISL_732540\|20 | ···················· | ···················· | ····················· | ···················· | ···················· | ···················· |
| Y | 4524 hCoV-19/Germany/BY-IMB00234/2020\|EPI_ISL_732541\|20 | ···················· | ···················· | ····················· | ···················· | ···················· | ···················· |
| Y | 4525 hCoV-19/Germany/BY-IMB01258/2020\|EPI_ISL_732542\|20 | ···················· | ···················· | ····················· | ···················· | ···················· | ···················· |
| Y | 4526 hCoV-19/Germany/BY-IMB01259/2020\|EPI_ISL_732543\|20 | ···················· | ···················· | ····················· | ···················· | ···················· | ···················· |
| Y | 4527 hCoV-19/Germany/BY-IMB01432/2020\|EPI_ISL_732544\|20 | ···················· | ···················· | ····················· | ···················· | ···················· | ···················· |
| Y | 4528 hCoV-19/Germany/BY-IMB02059/2020\|EPI_ISL_732545\|20 | ···················· | ···················· | ····················· | ···················· | ···················· | ···················· |
| Y | 4529 hCoV-19/Germany/BY-IMB02204/2020\|EPI_ISL_732546\|20 | ···················· | ···················· | ····················· | ···················· | ···················· | ···················· |
| Y | 4530 hCoV-19/Germany/BY-IMB02460/2020\|EPI_ISL_732547\|20 | ···················· | ···················· | ····················· | ···················· | ···················· | ···················· |
| Y | 4531 hCoV-19/Germany/BY-IMB02772/2020\|EPI_ISL_732548\|20 | ···················· | ···················· | ····················· | ···················· | ···················· | ···················· |
| Y | 4532 hCoV-19/Germany/BY-IMB02858/2020\|EPI_ISL_732549\|20 | ···················· | ···················· | ····················· | ···················· | ···················· | ···················· |
| Y | 4533 hCoV-19/Germany/BY-IMB02995/2020\|EPI_ISL_732550\|20 | ···················· | ···················· | ····················· | ···················· | ···················· | ···················· |
| Y | 4534 hCoV-19/Germany/BY-IMB03040/2020\|EPI_ISL_732551\|20 | ···················· | ···················· | ····················· | ···················· | ···················· | ···················· |
| Y | 4535 hCoV-19/Germany/BY-IMB03120/2020\|EPI_ISL_732552\|20 | ···················· | ···················· | ····················· | ···················· | ···················· | ···················· |
| Y | 4536 hCoV-19/Germany/BY-IMB03735/2020\|EPI_ISL_732553\|20 | ···················· | ···················· | ····················· | ···················· | ···················· | ···················· |
| Y | 4537 hCoV-19/Germany/BY-IMB04157/2020\|EPI_ISL_732554\|20 | ···················· | ···················· | ····················· | ···················· | ···················· | ···················· |
| Y | 4538 hCoV-19/Germany/BY-IMB04242/2020\|EPI_ISL_732555\|20 | ···················· | ···················· | ····················· | ···················· | ···················· | ···················· |
| Y | 4539 hCoV-19/Germany/BY-IMB05401/2020\|EPI_ISL_732556\|20 | ···················· | ···················· | ····················· | ···················· | ···················· | ···················· |
| Y | 4540 hCoV-19/Germany/BY-IMB06128/2020\|EPI_ISL_732557\|20 | ···················· | ···················· | ····················· | ···················· | ···················· | ···················· |
| Y | 4541 hCoV-19/Germany/BY-IMB06133/2020\|EPI_ISL_732558\|20 | ···················· | ···················· | ····················· | ···················· | ···················· | ···················· |
| Y | 4542 hCoV-19/Germany/BY-IMB06351/2020\|EPI_ISL_732559\|20 | ···················· | ···················· | ····················· | ···················· | ···················· | ···················· |
| Y | 4543 hCoV-19/Germany/BY-IMB06494/2020\|EPI_ISL_732560\|20 | ···················· | ···················· | ····················· | ···················· | ···················· | ···················· |
| Y | 4544 hCoV-19/Germany/BY-IMB11048/2020\|EPI_ISL_732561\|20 | ···················· | ·A·················· | ····················· | ···················· | ···················· | ···················· |
| Y | 4545 hCoV-19/Germany/BY-IMB11050/2020\|EPI_ISL_732562\|20 | ···················· | ·A·················· | ····················· | ···················· | ···················· | ···················· |
| Y | 4546 hCoV-19/Germany/BY-IMB11609/2020\|EPI_ISL_732563\|20 | ···················· | ···················· | ····················· | ···················· | ···················· | ···················· |
| Y | 4547 hCoV-19/Germany/NW-IMB01523/2020\|EPI_ISL_732564\|20 | ···················· | ···················· | ····················· | ···················· | ···················· | ···················· |
| Y | 4548 hCoV-19/Finland/11CA8/2020\|EPI_ISL_732565\|2020-10- | ···················· | ·A·················· | ····················· | ···················· | ···················· | ··A················· |
| Y | 4549 hCoV-19/Finland/11CB2/2020\|EPI_ISL_732566\|2020-10- | ···················· | ·A·················· | ····················· | ···················· | ···················· | ··A················· |
| Y | 4550 hCoV-19/Finland/11CD8/2020\|EPI_ISL_732568\|2020-10- | ···················· | ···················· | ····················· | ···················· | ···················· | ···················· |
| Y | 4551 hCoV-19/Finland/11CD9/2020\|EPI_ISL_732569\|2020-10- | ···················· | ···················· | ····················· | ···················· | ···················· | ···················· |
| Y | 4552 hCoV-19/Finland/11CE3/2020\|EPI_ISL_732570\|2020-12- | ···················· | ···················· | ····················· | ···················· | ···················· | ···················· |

| | | | | | | | |
|---|---|---|---|---|---|---|---|
| Y | 4553 hCoV-19/Finland/11CF3/2020\|EPI_ISL_732571\|2020-10- | ···················· | ·A·················· | ···················· | ···················· | ···················· | ··A················· |
| Y | 4554 hCoV-19/Finland/11CF4/2020\|EPI_ISL_732572\|2020-12- | ···················· | ···················· | ···················· | ···················· | ···················· | ···················· |
| Y | 4555 hCoV-19/Finland/11CH4/2020\|EPI_ISL_732573\|2020-10- | ···················· | ···················· | ···················· | ···················· | ···················· | ···················· |
| Y | 4556 hCoV-19/Finland/13CA6/2020\|EPI_ISL_732574\|2020-12- | ···················· | ···················· | ···················· | ···················· | ···················· | ···················· |
| Y | 4557 hCoV-19/Finland/13CB7/2020\|EPI_ISL_732575\|2020-12- | ···················· | ·A·················· | ···················· | ···················· | ···················· | ···················· |
| Y | 4558 hCoV-19/Finland/14CA7/2020\|EPI_ISL_732576\|2020-12- | ···················· | ···················· | ···················· | ···················· | ···················· | ···················· |
| Y | 4559 hCoV-19/Finland/14CA9/2020\|EPI_ISL_732577\|2020-12- | ···················· | ···················· | ···················· | ···················· | ···················· | ···················· |
| Y | 4560 hCoV-19/Finland/14CB12/2020\|EPI_ISL_732578\|2020-12 | ···················· | ···················· | ···················· | ···················· | ···················· | ···················· |
| Y | 4561 hCoV-19/Finland/14CB4/2020\|EPI_ISL_732579\|2020-12- | ···················· | ···················· | ···················· | ···················· | ···················· | ···················· |
| Y | 4562 hCoV-19/Finland/14CC10/2020\|EPI_ISL_732580\|2020-12 | ···················· | ···················· | ···················· | ···················· | ···················· | ···················· |
| Y | 4563 hCoV-19/Finland/14CC8/2020\|EPI_ISL_732581\|2020-12- | ···················· | ···················· | ···················· | ···················· | ···················· | ···················· |
| Y | 4564 hCoV-19/Finland/14CD8/2020\|EPI_ISL_732582\|2020-12- | ···················· | ···················· | ···················· | ···················· | ···················· | ···················· |
| Y | 4565 hCoV-19/Finland/14CG10/2020\|EPI_ISL_732583\|2020-12 | ···················· | ···················· | ···················· | ···················· | ···················· | ···················· |
| Y | 4566 hCoV-19/Finland/14CH8/2020\|EPI_ISL_732584\|2020-12- | ···················· | ·A·················· | ···················· | ···················· | ···················· | ···················· |
| Y | 4567 hCoV-19/Finland/15CE1/2020\|EPI_ISL_732585\|2020-12- | ···················· | ···················· | ···················· | ···················· | ···················· | ···················· |
| Y | 4568 hCoV-19/Finland/15CE5/2020\|EPI_ISL_732586\|2020-12- | ···················· | ···················· | ···················· | ···················· | ···················· | ···················· |
| Y | 4569 hCoV-19/Finland/16CC4/2020\|EPI_ISL_732587\|2020-12- | ···················· | ···················· | ···················· | ···················· | ···················· | ···················· |
| Y | 4570 hCoV-19/Finland/16CD2/2020\|EPI_ISL_732588\|2020-12- | ···················· | ·A·················· | ···················· | ···················· | ···················· | ··A················· |
| Y | 4571 hCoV-19/Finland/16CD4/2020\|EPI_ISL_732589\|2020-12- | ···················· | ···················· | ···················· | ···················· | ···················· | ···················· |
| Y | 4572 hCoV-19/Finland/16CD9/2020\|EPI_ISL_732590\|2020-12- | ···················· | ·A·················· | ···················· | ···················· | ···················· | ···················· |
| Y | 4573 hCoV-19/Finland/16CD9-b/2020\|EPI_ISL_732591\|2020-1 | ···················· | ···················· | ···················· | ···················· | ···················· | ···················· |
| Y | 4574 hCoV-19/Finland/16CF3/2020\|EPI_ISL_732592\|2020-12- | ···················· | ···················· | ···················· | ···················· | ···················· | ···················· |
| Y | 4575 hCoV-19/Finland/16CH7/2020\|EPI_ISL_732593\|2020-12- | ···················· | ···················· | ···················· | ···················· | ···················· | ···················· |
| Y | 4576 hCoV-19/Finland/18CE5/2020\|EPI_ISL_732594\|2020-12- | ···················· | ·A·················· | ···················· | ···················· | ···················· | ···················· |
| Y | 4577 hCoV-19/Finland/19CB11/2020\|EPI_ISL_732595\|2020-12 | ···················· | ···················· | ···················· | ···················· | ···················· | ···················· |
| Y | 4578 hCoV-19/Finland/19CG7/2020\|EPI_ISL_732596\|2020-12- | ···················· | ···················· | ···················· | ···················· | ···················· | ···················· |
| Y | 4579 hCoV-19/Finland/1CC1/2020\|EPI_ISL_732597\|2020-12-0 | ···················· | ···················· | ···················· | ···················· | ···················· | ···················· |
| Y | 4580 hCoV-19/Finland/1CD5/2020\|EPI_ISL_732598\|2020-12-0 | ···················· | ···················· | ···················· | ···················· | ···················· | ···················· |
| Y | 4581 hCoV-19/Finland/1cE11/2020\|EPI_ISL_732599\|2020-10- | ···················· | ·A·················· | ···················· | ···················· | ···················· | ···················· |
| Y | 4582 hCoV-19/England/CAMC-C2C8EC/2020\|EPI_ISL_733812\|20 | ···················· | ···················· | ···················· | ···················· | ···················· | ···················· |
| Y | 4583 hCoV-19/England/MILK-C2FBB3/2020\|EPI_ISL_733813\|20 | ···················· | ···················· | ···················· | ···················· | ···················· | ···················· |
| Y | 4584 hCoV-19/England/ALDP-C240CE/2020\|EPI_ISL_733815\|20 | ···················· | ···················· | ···················· | ···················· | ···················· | ···················· |
| Y | 4585 hCoV-19/England/ALDP-C240EC/2020\|EPI_ISL_733818\|20 | ···················· | ···················· | ···················· | ···················· | ···················· | ···················· |
| Y | 4586 hCoV-19/England/MILK-C2FAB6/2020\|EPI_ISL_733820\|20 | ···················· | ···················· | ···················· | ···················· | ···················· | ···················· |
| Y | 4587 hCoV-19/England/MILK-C2FB95/2020\|EPI_ISL_733828\|20 | ···················· | ···················· | ···················· | ···················· | ···················· | ···················· |
| Y | 4588 hCoV-19/England/MILK-C2F9B9/2020\|EPI_ISL_733829\|20 | ···················· | ···················· | ···················· | ···················· | ···················· | ···················· |
| Y | 4589 hCoV-19/England/MILK-C2FA6B/2020\|EPI_ISL_733832\|20 | ···················· | ···················· | ···················· | ···················· | ···················· | ···················· |
| Y | 4590 hCoV-19/England/ALDP-C23205/2020\|EPI_ISL_733833\|20 | ···················· | ···················· | ···················· | ···················· | ···················· | ···················· |
| Y | 4591 hCoV-19/England/CAMC-C2D520/2020\|EPI_ISL_733860\|20 | ···················· | ···················· | ···················· | ···················· | ···················· | ···················· |
| Y | 4592 hCoV-19/England/MILK-C2FA01/2020\|EPI_ISL_733861\|20 | ···················· | ···················· | ···················· | ···················· | ······T············· | ···················· |
| Y | 4593 hCoV-19/England/CAMC-C2D405/2020\|EPI_ISL_733862\|20 | ···················· | ···················· | ···················· | ···················· | ···················· | ···················· |
| Y | 4594 hCoV-19/England/ALDP-C23C9E/2020\|EPI_ISL_733867\|20 | ···················· | ···················· | ···················· | ···················· | ···················· | ···················· |
| Y | 4595 hCoV-19/England/CAMC-C26141/2020\|EPI_ISL_733868\|20 | ···················· | ···················· | ···················· | ···················· | ···················· | ···················· |
| Y | 4596 hCoV-19/England/MILK-C2FB2C/2020\|EPI_ISL_733872\|20 | ···················· | ···················· | ···················· | ···················· | ···················· | ···················· |
| Y | 4597 hCoV-19/England/ALDP-C23092/2020\|EPI_ISL_733877\|20 | ···················· | ···················· | ···················· | ···················· | ···················· | ···················· |
| Y | 4598 hCoV-19/England/ALDP-C22E7B/2020\|EPI_ISL_733880\|20 | ···················· | ···················· | ···················· | ···················· | ···················· | ···················· |
| Y | 4599 hCoV-19/England/ALDP-C23CCB/2020\|EPI_ISL_733882\|20 | ···················· | ···················· | ···················· | ···················· | ······T············· | ···················· |
| Y | 4600 hCoV-19/England/CAMC-C2CF50/2020\|EPI_ISL_733886\|20 | ···················· | ···················· | ···················· | ···················· | ···················· | ···················· |
| Y | 4601 hCoV-19/England/MILK-C2FFB7/2020\|EPI_ISL_733889\|20 | ···················· | ···················· | ···················· | ···················· | ···················· | ···················· |
| Y | 4602 hCoV-19/England/CAMC-C267B8/2020\|EPI_ISL_733892\|20 | ···················· | ···················· | ···················· | ···················· | ···················· | ···················· |
| Y | 4603 hCoV-19/England/ALDP-C22FB4/2020\|EPI_ISL_733897\|20 | ···················· | ···················· | ···················· | ···················· | ···················· | ···················· |
| Y | 4604 hCoV-19/England/QEUH-C5CB53/2020\|EPI_ISL_736090\|20 | ···················· | ···················· | ···················· | ···················· | ···················· | ···················· |
| Y | 4605 hCoV-19/England/ALDP-C57C55/2020\|EPI_ISL_736093\|20 | ···················· | ···················· | ···················· | ···················· | ···················· | ···················· |
| Y | 4606 hCoV-19/England/ALDP-C5888D/2020\|EPI_ISL_736095\|20 | ···················· | ···················· | ···················· | ···················· | ···················· | ···················· |
| Y | 4607 hCoV-19/England/ALDP-C588BA/2020\|EPI_ISL_736097\|20 | ···················· | ···················· | ···················· | ···················· | ···················· | ···················· |
| Y | 4608 hCoV-19/England/QEUH-C5BC8E/2020\|EPI_ISL_736099\|20 | ···················· | ···················· | ···················· | ···················· | ···················· | ···················· |
| Y | 4610 hCoV-19/England/NORW-F9D8B/2020\|EPI_ISL_725441\|202 | ···················· | ···················· | ···················· | ···················· | ···················· | ···················· |
| Y | 4611 hCoV-19/England/NORW-F9DE5/2020\|EPI_ISL_725443\|202 | ···················· | ···················· | ···················· | ···················· | ···················· | ···················· |
| Y | 4612 hCoV-19/England/NORW-F9EA6/2020\|EPI_ISL_725449\|202 | ···················· | ···················· | ···················· | ···················· | ···················· | ···················· |
| Y | 4613 hCoV-19/England/NORW-F9EE2/2020\|EPI_ISL_725450\|202 | ···················· | ···················· | ···················· | ···················· | ···················· | ···················· |
| Y | 4614 hCoV-19/England/NORW-F9F67/2020\|EPI_ISL_725455\|202 | ···················· | ···················· | ···················· | ···················· | ···················· | ···················· |
| Y | 4615 hCoV-19/England/NORW-FA0FA/2020\|EPI_ISL_725470\|202 | ···················· | ···················· | ···················· | ···················· | ···················· | ···················· |
| Y | 4616 hCoV-19/England/NORW-FA106/2020\|EPI_ISL_725471\|202 | ···················· | ···················· | ···················· | ···················· | ···················· | ···················· |
| Y | 4617 hCoV-19/England/NORW-FA124/2020\|EPI_ISL_725472\|202 | ···················· | ···················· | ···················· | ···················· | ···················· | ···················· |
| Y | 4618 hCoV-19/England/NORW-FA133/2020\|EPI_ISL_725473\|202 | ···················· | ···················· | ···················· | ···················· | ···················· | ···················· |
| Y | 4619 hCoV-19/England/NORW-FA17F/2020\|EPI_ISL_725477\|202 | ···················· | ···················· | ···················· | ···················· | ···················· | ···················· |
| Y | 4620 hCoV-19/England/NORW-FA1E8/2020\|EPI_ISL_725481\|202 | ···················· | ···················· | ···················· | ···················· | ···················· | ···················· |
| Y | 4621 hCoV-19/England/NORW-FA203/2020\|EPI_ISL_725482\|202 | ···················· | ···················· | ···················· | ···················· | ···················· | ···················· |
| Y | 4622 hCoV-19/England/NORW-FA230/2020\|EPI_ISL_725483\|202 | ···················· | ···················· | ···················· | ···················· | ···················· | ···················· |
| Y | 4623 hCoV-19/England/NORW-FA24F/2020\|EPI_ISL_725484\|202 | ···················· | ···················· | ···················· | ···················· | ···················· | ···················· |
| Y | 4624 hCoV-19/England/NORW-FA27C/2020\|EPI_ISL_725486\|202 | ···················· | ···················· | ···················· | ···················· | ···················· | ···················· |
| Y | 4625 hCoV-19/England/NORW-FA2B8/2020\|EPI_ISL_725489\|202 | ···················· | ···················· | ···················· | ···················· | ···················· | ···················· |
| Y | 4626 hCoV-19/England/NORW-FA2C7/2020\|EPI_ISL_725490\|202 | ···················· | ···················· | ···················· | ···················· | ···················· | ···················· |
| Y | 4627 hCoV-19/England/NORW-FA31F/2020\|EPI_ISL_725495\|202 | ···················· | ···················· | ···················· | ···················· | ···················· | ···················· |
| Y | 4629 hCoV-19/England/NORW-FA36A/2020\|EPI_ISL_725498\|202 | ···················· | ···················· | ···················· | ···················· | ···················· | ···················· |
| Y | 4630 hCoV-19/Wales/PHWC-497073/2020\|EPI_ISL_726771\|2020 | ···················· | ···················· | ···················· | ···················· | ···················· | ···················· |
| Y | 4631 hCoV-19/Wales/PHWC-497082/2020\|EPI_ISL_726772\|2020 | ···················· | ···················· | ···················· | ···················· | ···················· | ···················· |
| Y | 4639 hCoV-19/Wales/PHWC-497107/2020\|EPI_ISL_726780\|2020 | ···················· | ···················· | ···················· | ···················· | ···················· | ···················· |
| Y | 4640 hCoV-19/Wales/PHWC-497116/2020\|EPI_ISL_726781\|2020 | ···················· | ···················· | ···················· | ···················· | ···················· | ···················· |
| Y | 4641 hCoV-19/Wales/PHWC-497125/2020\|EPI_ISL_726782\|2020 | ···················· | ···················· | ···················· | ···················· | ···················· | ···················· |

| | | | | | | | | | |
|---|---|---|---|---|---|---|---|---|---|
| Y | 4643 | hCoV-19/Wales/PHWC-497143/2020\|EPI_ISL_726784\|2020 | ···················· | ···················· | ···················· | ···················· | ···················· | ···················· | |
| Y | 4644 | hCoV-19/Wales/PHWC-497152/2020\|EPI_ISL_726785\|2020 | ···················· | ···················· | ···················· | ···················· | ···················· | ···················· | |
| Y | 4645 | hCoV-19/Wales/PHWC-497161/2020\|EPI_ISL_726786\|2020 | ···················· | ···················· | ···················· | ···················· | ···················· | ···················· | |
| Y | 4646 | hCoV-19/Wales/PHWC-497170/2020\|EPI_ISL_726787\|2020 | ···················· | ···················· | ···················· | ···················· | ···················· | ···················· | |
| Y | 4647 | hCoV-19/Wales/PHWC-49718F/2020\|EPI_ISL_726788\|2020 | ···················· | ···················· | ···················· | ···················· | ···················· | ···················· | |
| Y | 4648 | hCoV-19/Wales/PHWC-49719E/2020\|EPI_ISL_726789\|2020 | ···················· | ···················· | ···················· | ···················· | ···················· | ···················· | |
| Y | 4649 | hCoV-19/Wales/PHWC-498E69/2020\|EPI_ISL_727220\|2020 | ···················· | ···················· | ···················· | ···················· | ···················· | ···················· | |
| Y | 4650 | hCoV-19/Wales/PHWC-498E87/2020\|EPI_ISL_727221\|2020 | ···················· | ···················· | ···················· | ···················· | ···················· | ···················· | |
| Y | 4651 | hCoV-19/Wales/PHWC-498ED2/2020\|EPI_ISL_727226\|2020 | ···················· | ···················· | ···················· | ···················· | ···················· | ···················· | |
| Y | 4655 | hCoV-19/Wales/PHWC-498F48/2020\|EPI_ISL_727232\|2020 | ···················· | ···················· | ···················· | ···················· | ···················· | ···················· | |
| Y | 4663 | hCoV-19/Wales/PHWC-498FEE/2020\|EPI_ISL_727242\|2020 | ···················· | ···················· | ···················· | ···················· | ···················· | ···················· | |
| Y | 4666 | hCoV-19/Wales/PHWC-499026/2020\|EPI_ISL_727246\|2020 | ···················· | ···················· | ···················· | ···················· | ···················· | ···················· | |
| Y | 4671 | hCoV-19/Wales/PHWC-49909F/2020\|EPI_ISL_727253\|2020 | ···················· | ···················· | ···················· | ···················· | ···················· | ···················· | |
| Y | 4675 | hCoV-19/Wales/PHWC-4990F9/2020\|EPI_ISL_727257\|2020 | ···················· | ······A············· | ···················· | ···················· | ···················· | ···················· | |
| Y | 4681 | hCoV-19/Wales/PHWC-4991AB/2020\|EPI_ISL_727267\|2020 | ···················· | ···················· | ···················· | ···················· | ···················· | ···················· | |
| Y | 4682 | hCoV-19/Wales/PHWC-4991BA/2020\|EPI_ISL_727268\|2020 | ···················· | ···················· | ···················· | ···················· | NN·················· | ···················· | |
| Y | 4684 | hCoV-19/Wales/PHWC-4991D8/2020\|EPI_ISL_727270\|2020 | ···················· | ···················· | ···················· | ···················· | ···················· | ···················· | |
| Y | 4685 | hCoV-19/Wales/PHWC-4991E7/2020\|EPI_ISL_727271\|2020 | ···················· | ···················· | ···················· | ···················· | ···················· | ···················· | |
| Y | 4688 | hCoV-19/Wales/PHWC-499220/2020\|EPI_ISL_727274\|2020 | ···················· | ···················· | ···················· | ···················· | ···················· | ···················· | |
| Y | 4689 | hCoV-19/Wales/PHWC-49923F/2020\|EPI_ISL_727275\|2020 | ···················· | ···················· | ···················· | ···················· | NNN················· | ···················· | 1 |
| Y | 4690 | hCoV-19/Wales/PHWC-49924E/2020\|EPI_ISL_727276\|2020 | ···················· | ···················· | ···················· | ···················· | ···················· | ···················· | |
| Y | 4693 | hCoV-19/Wales/PHWC-49927B/2020\|EPI_ISL_727279\|2020 | ···················· | ···················· | ···················· | ···················· | ···················· | ···················· | |
| Y | 4694 | hCoV-19/Wales/PHWC-49928A/2020\|EPI_ISL_727280\|2020 | ···················· | ···················· | ···················· | ···················· | ···················· | ···················· | |
| Y | 4695 | hCoV-19/Wales/PHWC-499299/2020\|EPI_ISL_727281\|2020 | ···················· | ···················· | ···················· | ···················· | ···················· | ···················· | |
| Y | 4697 | hCoV-19/Wales/PHWC-4992C6/2020\|EPI_ISL_727284\|2020 | ···················· | ···················· | ···················· | ···················· | ···················· | ···················· | |
| Y | 4698 | hCoV-19/Wales/PHWC-4992D5/2020\|EPI_ISL_727285\|2020 | ···················· | ···················· | ···················· | ···················· | ···················· | ···················· | |
| Y | 4699 | hCoV-19/Wales/PHWC-4992E4/2020\|EPI_ISL_727286\|2020 | ···T················ | ···················· | ···················· | ···················· | ···················· | ···················· | |
| Y | 4700 | hCoV-19/Wales/PHWC-4992F3/2020\|EPI_ISL_727287\|2020 | ···················· | ···················· | ···················· | ···················· | ···················· | ···················· | |
| Y | 4702 | hCoV-19/Wales/PHWC-49932D/2020\|EPI_ISL_727290\|2020 | ···················· | ···················· | ···················· | ···················· | ···················· | ···················· | |
| Y | 4706 | hCoV-19/Wales/PHWC-499378/2020\|EPI_ISL_727294\|2020 | ···················· | ···················· | ···················· | ···················· | ···················· | ···················· | |
| Y | 4707 | hCoV-19/Wales/PHWC-499387/2020\|EPI_ISL_727295\|2020 | ···················· | ···················· | ···················· | ···················· | ···················· | ···················· | |
| Y | 4710 | hCoV-19/Wales/PHWC-4993C3/2020\|EPI_ISL_727298\|2020 | ···················· | ···················· | ···················· | ···················· | ···················· | ···················· | |
| Y | 4712 | hCoV-19/France/ARA-111638/2020\|EPI_ISL_728550\|2020 | ···················· | ···················· | ···················· | ···················· | ···················· | ···················· | |
| Y | 4713 | hCoV-19/France/ARA-111718/2020\|EPI_ISL_728551\|2020 | ···················· | ···················· | ···················· | ···················· | ···················· | ···················· | |
| Y | 4714 | hCoV-19/Netherlands/UT-RIVM-20065/2020\|EPI_ISL_728 | ···················· | ···················· | ···················· | ···················· | ···················· | ···················· | |
| Y | 4715 | hCoV-19/Netherlands/NH-RIVM-20083/2020\|EPI_ISL_728 | ···················· | ···················· | ···················· | ···················· | ···················· | ···················· | |
| Y | 4716 | hCoV-19/Netherlands/NH-RIVM-20227/2020\|EPI_ISL_728 | ···················· | ···················· | ···················· | ···················· | ···················· | ···················· | |
| Y | 4717 | hCoV-19/Netherlands/NH-RIVM-20228/2020\|EPI_ISL_728 | ···················· | ···················· | ···················· | ···················· | ···················· | ···················· | |
| Y | 4718 | hCoV-19/Netherlands/NH-RIVM-20432/2020\|EPI_ISL_728 | ···················· | ···················· | ···················· | ···················· | ···················· | ···················· | |
| Y | 4719 | hCoV-19/Netherlands/ZH-RIVM-20349/2020\|EPI_ISL_728 | ···················· | ···················· | ···················· | ···················· | ···················· | ···················· | |
| Y | 4720 | hCoV-19/Netherlands/NB-RIVM-20350/2020\|EPI_ISL_728 | ···················· | ···················· | ···················· | ···················· | ···················· | ···················· | |
| Y | 4721 | hCoV-19/Netherlands/NB-RIVM-20351/2020\|EPI_ISL_728 | ···················· | ···················· | ···················· | ···················· | ···················· | ···················· | |
| Y | 4722 | hCoV-19/Netherlands/NB-RIVM-20352/2020\|EPI_ISL_728 | ···················· | ···················· | ···················· | ···················· | ···················· | ···················· | |
| Y | 4723 | hCoV-19/Netherlands/NB-RIVM-20353/2020\|EPI_ISL_728 | ···················· | ···················· | ···················· | ···················· | ···················· | ···················· | |
| Y | 4724 | hCoV-19/Netherlands/NB-RIVM-20354/2020\|EPI_ISL_728 | ···················· | ···················· | ···················· | ···················· | ···················· | ···················· | |
| Y | 4725 | hCoV-19/Netherlands/NB-RIVM-20355/2020\|EPI_ISL_728 | ···················· | ···················· | ···················· | ···················· | ···················· | ···················· | |
| Y | 4726 | hCoV-19/Netherlands/NB-RIVM-20356/2020\|EPI_ISL_728 | ···················· | ···················· | ···················· | ···················· | ······T············· | ···················· | |
| Y | 4727 | hCoV-19/Netherlands/UT-RIVM-20357/2020\|EPI_ISL_728 | ···················· | ···················· | ···················· | ···················· | ···················· | ···················· | |
| Y | 4728 | hCoV-19/Netherlands/UT-RIVM-20358/2020\|EPI_ISL_728 | ···················· | ···················· | ···················· | ···················· | ···················· | ···················· | |
| Y | 4729 | hCoV-19/Netherlands/UT-RIVM-20359/2020\|EPI_ISL_728 | ···················· | ···················· | ···················· | ···················· | ···················· | ···················· | |
| Y | 4730 | hCoV-19/Netherlands/FR-RIVM-20360/2020\|EPI_ISL_728 | ···················· | ···················· | ···················· | ···················· | ···················· | ···················· | |
| Y | 4731 | hCoV-19/Netherlands/FR-RIVM-20361/2020\|EPI_ISL_728 | ···················· | ···················· | ···················· | ···················· | ···················· | ···················· | |
| Y | 4732 | hCoV-19/Netherlands/FR-RIVM-20362/2020\|EPI_ISL_728 | ···················· | ···················· | ···················· | ···················· | ···················· | ···················· | |
| Y | 4733 | hCoV-19/Netherlands/UT-RIVM-20363/2020\|EPI_ISL_728 | ···················· | ···················· | ···················· | ···················· | ···················· | ···················· | |
| Y | 4734 | hCoV-19/Netherlands/UT-RIVM-20364/2020\|EPI_ISL_728 | ···················· | ···················· | ···················· | ···················· | ···················· | ···················· | |
| Y | 4735 | hCoV-19/Netherlands/ZH-RIVM-20365/2020\|EPI_ISL_728 | ···················· | ···················· | ···················· | ···················· | ···················· | ···················· | |
| Y | 4736 | hCoV-19/Netherlands/ZH-RIVM-20366/2020\|EPI_ISL_728 | ···················· | ···················· | ···················· | ···················· | ···················· | ···················· | |
| Y | 4737 | hCoV-19/Netherlands/ZH-RIVM-20367/2020\|EPI_ISL_728 | ···················· | ···················· | ···················· | ···················· | ···················· | ···················· | |
| Y | 4738 | hCoV-19/Netherlands/NB-RIVM-20368/2020\|EPI_ISL_728 | ···················· | ···················· | ···················· | ···················· | ···················· | ···················· | |
| Y | 4739 | hCoV-19/Netherlands/UT-RIVM-20369/2020\|EPI_ISL_728 | ···················· | ···················· | ···················· | ···················· | ···················· | ···················· | |
| Y | 4740 | hCoV-19/Netherlands/FR-RIVM-20370/2020\|EPI_ISL_728 | ···················· | ···················· | ···················· | ···················· | ···················· | ···················· | |
| Y | 4741 | hCoV-19/Netherlands/FR-RIVM-20371/2020\|EPI_ISL_728 | ···················· | ···················· | ···················· | ···················· | ···················· | ···················· | |
| Y | 4742 | hCoV-19/Netherlands/ZH-RIVM-20372/2020\|EPI_ISL_728 | ···················· | ···················· | ···················· | ···················· | ···················· | ···················· | |
| Y | 4743 | hCoV-19/Netherlands/UT-RIVM-20373/2020\|EPI_ISL_728 | ···················· | ···················· | ···················· | ···················· | ···················· | ···················· | |
| Y | 4744 | hCoV-19/Netherlands/ZH-RIVM-20374/2020\|EPI_ISL_728 | ···················· | ···················· | ···················· | ···················· | ···················· | ···················· | |
| Y | 4745 | hCoV-19/Netherlands/NB-RIVM-20375/2020\|EPI_ISL_728 | ···················· | ···················· | ···················· | ···················· | ···················· | ···················· | |
| Y | 4746 | hCoV-19/Netherlands/NB-RIVM-20376/2020\|EPI_ISL_728 | ···················· | ···················· | ···················· | ···················· | ···················· | ···················· | |
| Y | 4747 | hCoV-19/Netherlands/UT-RIVM-20377/2020\|EPI_ISL_728 | ···················· | ···················· | ···················· | ···················· | ···················· | ···················· | |
| Y | 4748 | hCoV-19/Netherlands/UT-RIVM-20378/2020\|EPI_ISL_728 | ···················· | ···················· | ···················· | ···················· | ···················· | ···················· | |
| Y | 4749 | hCoV-19/Netherlands/UT-RIVM-20379/2020\|EPI_ISL_728 | ···················· | ···················· | ···················· | ···················· | ···················· | ···················· | |
| Y | 4750 | hCoV-19/Peru/LIM-UPCH-0175/2020\|EPI_ISL_729880\|202 | ···················· | ···················· | ···················· | ···················· | ···················· | ···················· | |
| Y | 4751 | hCoV-19/Peru/LIM-UPCH-0176/2020\|EPI_ISL_729881\|202 | ···················· | ···················· | ···················· | ···················· | ···················· | ···················· | |
| Y | 4752 | hCoV-19/Peru/LIM-UPCH-0179/2020\|EPI_ISL_729882\|202 | ···················· | ···················· | ···················· | ···················· | ···················· | ···················· | |
| Y | 4753 | hCoV-19/Peru/LIM-UPCH-0182/2020\|EPI_ISL_729883\|202 | ···················· | ···················· | ···················· | ···················· | ···················· | ···················· | |
| Y | 4754 | hCoV-19/Peru/LIM-UPCH-0184/2020\|EPI_ISL_729884\|202 | ···················· | ···················· | ···················· | ···················· | ···················· | ···················· | |
| Y | 4755 | hCoV-19/Peru/LIM-UPCH-0187/2020\|EPI_ISL_729885\|202 | ···················· | ···················· | ···················· | ···················· | ···················· | ···················· | |
| Y | 4756 | hCoV-19/Peru/LIM-UPCH-0189/2020\|EPI_ISL_729886\|202 | ···················· | ···················· | ···················· | ···················· | ···················· | ···················· | |
| Y | 4757 | hCoV-19/Peru/LIM-UPCH-0191/2020\|EPI_ISL_729887\|202 | ···················· | ···················· | ···················· | ···················· | ···················· | ···················· | |
| Y | 4758 | hCoV-19/Peru/LIM-UPCH-0193/2020\|EPI_ISL_729888\|202 | ···················· | ···················· | ···················· | ···················· | ···················· | ···················· | |
| Y | 4759 | hCoV-19/Peru/LIM-UPCH-0194/2020\|EPI_ISL_729889\|202 | ···················· | ···················· | ···················· | ···················· | ···················· | ···················· | |

| | | | | | | | | |
|---|---|---|---|---|---|---|---|---|
| Y | 4760 hCoV-19/Peru/LIM-UPCH-0196/2020\|EPI_ISL_729890\|202 | ···················· | ···················· | ···················· | ···················· | ···················· | ···················· | |
| Y | 4761 hCoV-19/Peru/LIM-UPCH-0197/2020\|EPI_ISL_729891\|202 | ···················· | ···················· | ···················· | ···················· | ···················· | ···················· | |
| Y | 4762 hCoV-19/Peru/LIM-UPCH-0198/2020\|EPI_ISL_729892\|202 | ···················· | ···················· | ···················· | ···················· | ···················· | ···················· | |
| Y | 4763 hCoV-19/Peru/LIM-UPCH-0200/2020\|EPI_ISL_729893\|202 | ···················· | ···················· | ···················· | ···················· | ···················· | ···················· | |
| Y | 4764 hCoV-19/Peru/LIM-UPCH-0202/2020\|EPI_ISL_729894\|202 | ···················· | ···················· | ···················· | ···················· | ···················· | ···················· | |
| Y | 4765 hCoV-19/Peru/LIM-UPCH-0204/2020\|EPI_ISL_729895\|202 | ···················· | ···················· | ···················· | ···················· | ···················· | ···················· | |
| Y | 4766 hCoV-19/Peru/LIM-UPCH-0205/2020\|EPI_ISL_729896\|202 | ···················· | ···················· | ···················· | ···················· | ···················· | ···················· | |
| Y | 4767 hCoV-19/Peru/LIM-UPCH-0207/2020\|EPI_ISL_729897\|202 | ···················· | ···················· | ···················· | ···················· | ···················· | ···················· | |
| Y | 4768 hCoV-19/Peru/LIM-UPCH-0236/2020\|EPI_ISL_729898\|202 | ···················· | ···················· | ···················· | ···················· | ···················· | ···················· | |
| Y | 4769 hCoV-19/Peru/LIM-UPCH-0240/2020\|EPI_ISL_729899\|202 | ···················· | ···················· | ···················· | ···················· | ···················· | ···················· | |
| Y | 4770 hCoV-19/England/QEUH-C31495/2020\|EPI_ISL_730864\|20 | ···················· | ···················· | ···················· | ···················· | ···················· | ···················· | |
| Y | 4771 hCoV-19/England/ALDP-C3D7FA/2020\|EPI_ISL_730865\|20 | ···················· | ···················· | ···················· | ···················· | ······T············· | ···················· | |
| Y | 4772 hCoV-19/England/QEUH-C317F6/2020\|EPI_ISL_730867\|20 | ···················· | ···················· | ···················· | ···················· | ······T············· | ···················· | |
| Y | 4773 hCoV-19/England/ALDP-C398DD/2020\|EPI_ISL_730868\|20 | ···················· | ···················· | ···················· | ···················· | ···················· | ···················· | |
| Y | 4774 hCoV-19/England/QEUH-C30FC2/2020\|EPI_ISL_730869\|20 | ···················· | ···················· | ···················· | ···················· | ···················· | ···················· | |
| Y | 4775 hCoV-19/England/ALDP-C3AEF8/2020\|EPI_ISL_730872\|20 | ···················· | ···················· | ···················· | ···················· | ···················· | ···················· | |
| Y | 4776 hCoV-19/England/ALDP-C3D61B/2020\|EPI_ISL_730873\|20 | ···················· | ···················· | ···················· | ···················· | ···················· | ···················· | |
| Y | 4777 hCoV-19/England/ALDP-C3B54D/2020\|EPI_ISL_730874\|20 | ···················· | ···················· | ···················· | ···················· | ···················· | ···················· | |
| Y | 4778 hCoV-19/England/QEUH-C317D8/2020\|EPI_ISL_730875\|20 | ···················· | ···················· | ···················· | ···················· | ···················· | ···················· | |
| Y | 4779 hCoV-19/England/ALDP-C3DEF5/2020\|EPI_ISL_730878\|20 | ···················· | ···················· | ···················· | ···················· | ···················· | ···················· | |
| Y | 4780 hCoV-19/England/ALDP-C3A5B7/2020\|EPI_ISL_730883\|20 | ···················· | ···················· | ···················· | ···················· | ···················· | ···················· | |
| Y | 4781 hCoV-19/England/ALDP-C39D74/2020\|EPI_ISL_730884\|20 | ···················· | ···················· | ···················· | ···················· | ···················· | ···················· | |
| Y | 4782 hCoV-19/England/ALDP-C3B668/2020\|EPI_ISL_730885\|20 | ···················· | ···················· | ···················· | ···················· | ···················· | ···················· | |
| Y | 4783 hCoV-19/England/ALDP-C3DBC1/2020\|EPI_ISL_730886\|20 | ···················· | ···················· | ···················· | ···················· | ···················· | ···················· | |
| Y | 4784 hCoV-19/England/QEUH-C317E7/2020\|EPI_ISL_730887\|20 | ···················· | ···················· | ···················· | ···················· | ···················· | ···················· | |
| Y | 4785 hCoV-19/England/ALDP-C3D9F4/2020\|EPI_ISL_730890\|20 | ···················· | ···················· | ···················· | ···················· | ···················· | ···················· | |
| Y | 4786 hCoV-19/England/ALDP-C3DEC8/2020\|EPI_ISL_730893\|20 | ···················· | ···················· | ···················· | ···················· | ···················· | ···················· | |
| Y | 4787 hCoV-19/England/QEUH-C317AB/2020\|EPI_ISL_730894\|20 | ···················· | ···················· | ···················· | ···················· | ···················· | ···················· | |
| Y | 4788 hCoV-19/England/ALDP-C3B57A/2020\|EPI_ISL_730895\|20 | ···················· | ···················· | ···················· | ···················· | ···················· | ···················· | |
| Y | 4789 hCoV-19/England/ALDP-C39D1A/2020\|EPI_ISL_730896\|20 | ···················· | ···················· | ···················· | ···················· | ···················· | ···················· | |
| Y | 4790 hCoV-19/USA/NY-Wadsworth-252369-01/2020\|EPI_ISL_73 | ···················· | ···················· | ···················· | ···················· | ···················· | ···················· | |
| Y | 4791 hCoV-19/USA/NY-Wadsworth-253001-01/2020\|EPI_ISL_73 | ···················· | ···················· | ···················· | ···················· | ···················· | ···················· | |
| Y | 4792 hCoV-19/USA/NY-Wadsworth-253061-01/2020\|EPI_ISL_73 | ···················· | ···················· | ···················· | ···················· | ···················· | ···················· | |
| Y | 4793 hCoV-19/USA/NY-Wadsworth-253063-01/2020\|EPI_ISL_73 | ···················· | ···················· | ···················· | ···················· | ···················· | ···················· | |
| Y | 4794 hCoV-19/USA/NY-Wadsworth-253441-01/2020\|EPI_ISL_73 | ···················· | ·A·················· | ···················· | ···················· | ···················· | ···················· | |
| Y | 4795 hCoV-19/USA/NY-Wadsworth-253622-01/2020\|EPI_ISL_73 | ···················· | ···················· | ···················· | ···················· | ···················· | ···················· | |
| Y | 4796 hCoV-19/USA/NY-Wadsworth-255287-01/2020\|EPI_ISL_73 | ···················· | ·A·················· | ···················· | ···················· | ···················· | ···················· | |
| Y | 4797 hCoV-19/USA/NY-Wadsworth-255625-01/2020\|EPI_ISL_73 | ···················· | ···················· | ···················· | ···················· | ···················· | ···················· | |
| Y | 4798 hCoV-19/USA/NY-Wadsworth-255794-01/2020\|EPI_ISL_73 | ···················· | ·A·················· | ···················· | ···················· | ···················· | ···················· | |
| Y | 4799 hCoV-19/USA/NY-Wadsworth-257969-01/2020\|EPI_ISL_73 | ···················· | ·············A······ | ···················· | ···················· | ···················· | ···················· | |
| Y | 4800 hCoV-19/USA/NY-Wadsworth-258235-01/2020\|EPI_ISL_73 | ···················· | ·A·················· | ···················· | ···················· | ···················· | ···················· | |
| Y | 4801 hCoV-19/USA/NY-Wadsworth-258524-01/2020\|EPI_ISL_73 | ···················· | ···················· | ···················· | ···················· | ···················· | ···················· | |
| Y | 4802 hCoV-19/USA/NY-Wadsworth-258525-01/2020\|EPI_ISL_73 | ···················· | ···················· | ···················· | ···················· | ···················· | ···················· | |
| Y | 4803 hCoV-19/USA/NY-Wadsworth-258600-01/2020\|EPI_ISL_73 | ···················· | ···················· | ···················· | ···················· | ···················· | ···················· | |
| Y | 4804 hCoV-19/USA/NY-Wadsworth-262689-01/2020\|EPI_ISL_73 | ···················· | ···················· | ···················· | ···················· | ···················· | ···················· | |
| Y | 4805 hCoV-19/USA/NY-Wadsworth-263158-01/2020\|EPI_ISL_73 | ···················· | ···················· | ···················· | ···················· | ···················· | ···················· | |
| Y | 4806 hCoV-19/USA/NY-Wadsworth-263161-01/2020\|EPI_ISL_73 | ···················· | ···················· | ···················· | ···················· | ···················· | ···················· | |
| Y | 4807 hCoV-19/USA/NY-Wadsworth-263168-01/2020\|EPI_ISL_73 | ···················· | ···················· | ···················· | ···················· | ···················· | ···················· | |
| Y | 4808 hCoV-19/USA/NY-Wadsworth-263180-01/2020\|EPI_ISL_73 | ···················· | ···················· | ···················· | ···················· | ···················· | ···················· | |
| Y | 4809 hCoV-19/USA/NY-Wadsworth-264178-01/2020\|EPI_ISL_73 | ···················· | ·A·················· | ···················· | ···················· | ···················· | ···················· | |
| Y | 4810 hCoV-19/USA/NY-Wadsworth-264179-01/2020\|EPI_ISL_73 | ···················· | ·A·················· | ···················· | ···················· | ···················· | ···················· | |
| Y | 4811 hCoV-19/USA/NY-Wadsworth-264183-01/2020\|EPI_ISL_73 | ···················· | ·A·················· | ···················· | ···················· | ···················· | ···················· | |
| Y | 4812 hCoV-19/USA/NY-Wadsworth-264185-01/2020\|EPI_ISL_73 | ···················· | ·A·················· | ···················· | ···················· | ···················· | ···················· | |
| Y | 4813 hCoV-19/USA/NY-Wadsworth-264187-01/2020\|EPI_ISL_73 | ···················· | ·A·················· | ···················· | ···················· | ···················· | ···················· | |
| Y | 4814 hCoV-19/USA/NY-Wadsworth-264198-01/2020\|EPI_ISL_73 | ···················· | ·A·················· | ···················· | ···················· | ···················· | ···················· | |
| Y | 4815 hCoV-19/USA/NY-Wadsworth-264209-01/2020\|EPI_ISL_73 | ···················· | ·A·················· | ···················· | ···················· | ···················· | ···················· | |
| Y | 4816 hCoV-19/USA/NY-Wadsworth-264213-01/2020\|EPI_ISL_73 | ···················· | ·A·················· | ···················· | ···················· | ···················· | ···················· | |
| Y | 4817 hCoV-19/USA/NY-Wadsworth-269589-01/2020\|EPI_ISL_73 | ···················· | ·A·················· | ···················· | ···················· | ···················· | ···················· | |
| Y | 4818 hCoV-19/England/MILK-C617C0/2020\|EPI_ISL_735701\|20 | ···················· | ···················· | ···················· | ···················· | ···················· | ···················· | |
| Y | 4819 hCoV-19/England/MILK-C6155D/2020\|EPI_ISL_735711\|20 | ···················· | ···················· | ···················· | ···················· | ···················· | ···················· | |
| Y | 4820 hCoV-19/England/MILK-C6220A/2020\|EPI_ISL_735713\|20 | ···················· | ···················· | ···················· | ···················· | ···················· | ···················· | |
| Y | 4821 hCoV-19/England/MILK-C6213A/2020\|EPI_ISL_735726\|20 | ···················· | ·A·················· | ···················· | ···················· | ···················· | ···················· | |
| Y | 4822 hCoV-19/England/MILK-C61A9A/2020\|EPI_ISL_735729\|20 | ···················· | ···················· | ···················· | ···················· | ···················· | ···················· | |
| Y | 4823 hCoV-19/England/MILK-C63931/2020\|EPI_ISL_735736\|20 | ···················· | ···················· | ···················· | ···················· | ···················· | ···················· | |
| Y | 4824 hCoV-19/England/MILK-C614F6/2020\|EPI_ISL_735737\|20 | ···················· | ···················· | ···················· | ···················· | ···················· | ···················· | |
| Y | 4825 hCoV-19/England/MILK-C638BC/2020\|EPI_ISL_735741\|20 | ···················· | ···················· | ···················· | ···················· | ···················· | ···················· | |
| Y | 4826 hCoV-19/England/MILK-C620A6/2020\|EPI_ISL_735742\|20 | ···················· | ···················· | ···················· | ···················· | ···················· | ···················· | |
| Y | 4827 hCoV-19/England/MILK-C62B1E/2020\|EPI_ISL_735743\|20 | ···················· | ···················· | ···················· | ···················· | ···················· | ···················· | |
| Y | 4828 hCoV-19/England/MILK-C63AB6/2020\|EPI_ISL_735745\|20 | ···················· | ···················· | ···················· | ···················· | ···················· | ···················· | |
| Y | 4829 hCoV-19/England/MILK-C61AC7/2020\|EPI_ISL_735753\|20 | ···················· | ···················· | ···················· | ···················· | ···················· | ···················· | |
| Y | 4830 hCoV-19/England/MILK-C624B9/2020\|EPI_ISL_735754\|20 | ··················T· | ···················· | ···················· | ···················· | ···················· | ···················· | 1 |
| Y | 4831 hCoV-19/England/MILK-C61E9E/2020\|EPI_ISL_735756\|20 | ···················· | ···················· | ···················· | ···················· | ······T············· | ···················· | |
| Y | 4832 hCoV-19/England/MILK-C61CD0/2020\|EPI_ISL_735758\|20 | ···················· | ···················· | ···················· | ···················· | ···················· | ···················· | |
| Y | 4833 hCoV-19/England/QEUH-C5CB26/2020\|EPI_ISL_736101\|20 | ···················· | ···················· | ···················· | ···················· | ···················· | ···················· | |
| Y | 4834 hCoV-19/England/ALDP-C582CB/2020\|EPI_ISL_736103\|20 | ···················· | ···················· | ···················· | ···················· | ···················· | ···················· | |
| Y | 4835 hCoV-19/England/ALDP-C58717/2020\|EPI_ISL_736106\|20 | ···················· | ···················· | ···················· | ···················· | ···················· | ···················· | |
| Y | 4836 hCoV-19/England/QEUH-C5D4B1/2020\|EPI_ISL_736107\|20 | ···················· | ···················· | ···················· | ···················· | ···················· | ···················· | |
| Y | 4837 hCoV-19/England/ALDP-C588D8/2020\|EPI_ISL_736108\|20 | ···················· | ···················· | ···················· | ···················· | ···················· | ···················· | |
| Y | 4838 hCoV-19/England/ALDP-C58832/2020\|EPI_ISL_736111\|20 | ···················· | ···················· | ···················· | ···················· | ······T············· | ···················· | |
| Y | 4839 hCoV-19/England/ALDP-C591BE/2020\|EPI_ISL_736117\|20 | ···················· | ···················· | ···················· | ···················· | ···················· | ···················· | |

| | | | | | | | | |
|---|---|---|---|---|---|---|---|---|
| Y | 4840 | hCoV-19/England/ALDP-C57D52/2020\|EPI_ISL_736118\|20 | ···················· | ···················· | ···················· | ···················· | ···················· | ···················· |
| Y | 4841 | hCoV-19/England/ALDP-C591CD/2020\|EPI_ISL_736119\|20 | ···················· | ···················· | ···················· | ···················· | ···················· | ···················· |
| Y | 4842 | hCoV-19/England/ALDP-C580C1/2020\|EPI_ISL_736122\|20 | ···················· | ···················· | ···················· | ···················· | ···················· | ···················· |
| Y | 4843 | hCoV-19/England/ALDP-C58261/2020\|EPI_ISL_736123\|20 | ···················· | ···················· | ···················· | ···················· | ···················· | ···················· |
| Y | 4844 | hCoV-19/England/ALDP-C5850E/2020\|EPI_ISL_736125\|20 | ···················· | ···················· | ···················· | ···················· | ···················· | ···················· |
| Y | 4845 | hCoV-19/England/ALDP-C57F7A/2020\|EPI_ISL_736127\|20 | ···················· | ···················· | ···················· | ···················· | ···················· | ···················· |
| Y | 4846 | hCoV-19/England/ALDP-C597AD/2020\|EPI_ISL_736128\|20 | ···················· | ···················· | ···················· | ···················· | ···················· | ···················· |
| Y | 4847 | hCoV-19/England/QEUH-C5BD21/2020\|EPI_ISL_736130\|20 | ···················· | ·········C·········· | ···················· | ···················· | ···················· | ···················· |
| Y | 4848 | hCoV-19/England/QEUH-C5D475/2020\|EPI_ISL_736131\|20 | ···················· | ···················· | ···················· | ···················· | ···················· | ···················· |
| Y | 4849 | hCoV-19/England/QEUH-C5D572/2020\|EPI_ISL_736132\|20 | ···················· | ···················· | ···················· | ···················· | ···················· | ···················· |
| Y | 4850 | hCoV-19/England/ALDP-C57F6B/2020\|EPI_ISL_736134\|20 | ···················· | ···················· | ···················· | ···················· | ···················· | ···················· |
| Y | 4851 | hCoV-19/England/ALDP-C591DC/2020\|EPI_ISL_736136\|20 | ···················· | ···················· | ···················· | ···················· | ···················· | ···················· |
| Y | 4852 | hCoV-19/England/ALDP-C58270/2020\|EPI_ISL_736140\|20 | ···················· | ···················· | ···················· | ···················· | ···················· | ···················· |
| Y | 4853 | hCoV-19/England/ALDP-C58C27/2020\|EPI_ISL_736141\|20 | ···················· | ···················· | ···················· | ···················· | ···················· | ···················· |
| Y | 4854 | hCoV-19/England/ALDP-C57F98/2020\|EPI_ISL_736142\|20 | ···················· | ···················· | ···················· | ···················· | ···················· | ···················· |
| Y | 4855 | hCoV-19/England/ALDP-C582BC/2020\|EPI_ISL_736144\|20 | ···················· | ···················· | ···················· | ···················· | ···················· | ···················· |
| Y | 4856 | hCoV-19/England/ALDP-C5978F/2020\|EPI_ISL_736148\|20 | ···················· | ···················· | ···················· | ···················· | ···················· | ···················· |
| Y | 4857 | hCoV-19/England/ALDP-C58A5A/2020\|EPI_ISL_736150\|20 | ···················· | ···················· | ···················· | ···················· | ···················· | ···················· |
| Y | 4858 | hCoV-19/England/ALDP-C5803A/2020\|EPI_ISL_736152\|20 | ···················· | ···················· | ···················· | ···················· | ···················· | ···················· |
| Y | 4859 | hCoV-19/England/ALDP-C57F5C/2020\|EPI_ISL_736153\|20 | ···················· | ···················· | ···················· | ···················· | ···················· | ···A················ |
| Y | 4860 | hCoV-19/England/ALDP-C57C19/2020\|EPI_ISL_736158\|20 | ···················· | ···················· | ···················· | ···················· | ···················· | ···················· |
| Y | 4861 | hCoV-19/England/QEUH-C5B78D/2020\|EPI_ISL_736162\|20 | ···················· | ···················· | ···················· | ···················· | ···················· | ···················· |
| Y | 4862 | hCoV-19/England/ALDP-C586FC/2020\|EPI_ISL_736164\|20 | ···················· | ···················· | ···················· | ···················· | ···················· | ···················· |
| Y | 4863 | hCoV-19/England/ALDP-C58164/2020\|EPI_ISL_736165\|20 | ···················· | ···················· | ···················· | ···················· | ···················· | ···················· |
| Y | 4864 | hCoV-19/England/QEUH-C5BCAC/2020\|EPI_ISL_736168\|20 | ···················· | ···················· | ···················· | ···················· | ···················· | ···················· |
| Y | 4866 | hCoV-19/Wales/PHWC-499554/2020\|EPI_ISL_727321\|2020 | ···················· | ···················· | ···················· | ···················· | ···················· | ···················· |
| Y | 4868 | hCoV-19/Wales/PHWC-499590/2020\|EPI_ISL_727324\|2020 | ···················· | ···················· | ···················· | ···················· | ···················· | ···················· |
| Y | 4869 | hCoV-19/Wales/PHWC-4995AF/2020\|EPI_ISL_727325\|2020 | ···················· | ···················· | ···················· | ···················· | ···················· | ···················· |
| Y | 4870 | hCoV-19/Wales/PHWC-4995BE/2020\|EPI_ISL_727326\|2020 | ···················· | ···················· | ···················· | ···················· | ···················· | ···················· |
| Y | 4871 | hCoV-19/Wales/PHWC-4995CD/2020\|EPI_ISL_727327\|2020 | ···················· | ···················· | ···················· | ···················· | ···················· | ···················· |
| Y | 4873 | hCoV-19/Wales/PHWC-4995EB/2020\|EPI_ISL_727329\|2020 | ···················· | ···················· | ···················· | ···················· | ···················· | ···················· |
| Y | 4874 | hCoV-19/Netherlands/UT-RIVM-20380/2020\|EPI_ISL_728 | ···················· | ···················· | ···················· | ···················· | ···················· | ···················· |
| Y | 4875 | hCoV-19/Netherlands/UT-RIVM-20381/2020\|EPI_ISL_728 | ···················· | ···················· | ···················· | ···················· | ···················· | ···················· |
| Y | 4876 | hCoV-19/Netherlands/UT-RIVM-20382/2020\|EPI_ISL_728 | ···················· | ···················· | ···················· | ···················· | ···················· | ···················· |
| Y | 4877 | hCoV-19/Netherlands/UT-RIVM-20383/2020\|EPI_ISL_728 | ···················· | ···················· | ···················· | ···················· | ···················· | ···················· |
| Y | 4878 | hCoV-19/Netherlands/FR-RIVM-20384/2020\|EPI_ISL_728 | ···················· | ···················· | ···················· | ···················· | ······T············· | ···················· |
| Y | 4879 | hCoV-19/Netherlands/FR-RIVM-20385/2020\|EPI_ISL_728 | ···················· | ···················· | ···················· | ···················· | ······T············· | ···················· |
| Y | 4880 | hCoV-19/Netherlands/FR-RIVM-20386/2020\|EPI_ISL_728 | ···················· | ···················· | ···················· | ···················· | ···················· | ···················· |
| Y | 4881 | hCoV-19/Netherlands/FR-RIVM-20387/2020\|EPI_ISL_728 | ···················· | ···················· | ···················· | ···················· | ···················· | ···················· |
| Y | 4882 | hCoV-19/Netherlands/FR-RIVM-20388/2020\|EPI_ISL_728 | ···················· | ···················· | ···················· | ···················· | ···················· | ···················· |
| Y | 4883 | hCoV-19/Netherlands/FR-RIVM-20389/2020\|EPI_ISL_728 | ···················· | ···················· | ···················· | ···················· | ···················· | ···················· |
| Y | 4884 | hCoV-19/Netherlands/ZH-RIVM-20390/2020\|EPI_ISL_728 | ···················· | ···················· | ···················· | ···················· | ···················· | ···················· |
| Y | 4885 | hCoV-19/Netherlands/UT-RIVM-20391/2020\|EPI_ISL_728 | ···················· | ···················· | ···················· | ···················· | ···················· | ···················· |
| Y | 4886 | hCoV-19/Netherlands/UT-RIVM-20392/2020\|EPI_ISL_728 | ···················· | ···················· | ···················· | ···················· | ···················· | ···················· |
| Y | 4887 | hCoV-19/Netherlands/UT-RIVM-20393/2020\|EPI_ISL_728 | ···················· | ···················· | ···················· | ···················· | ···················· | ···················· |
| Y | 4888 | hCoV-19/Netherlands/ZH-RIVM-20394/2020\|EPI_ISL_728 | ···················· | ···················· | ···················· | ···················· | ···················· | ···················· |
| Y | 4889 | hCoV-19/Netherlands/ZH-RIVM-20395/2020\|EPI_ISL_728 | ···················· | ···················· | ···················· | ···················· | ···················· | ···················· |
| Y | 4890 | hCoV-19/Netherlands/ZH-RIVM-20396/2020\|EPI_ISL_728 | ···················· | ···················· | ···················· | ···················· | ···················· | ···················· |
| Y | 4891 | hCoV-19/Netherlands/ZH-RIVM-20397/2020\|EPI_ISL_728 | ···················· | ···················· | ···················· | ···················· | ···················· | ···················· |
| Y | 4892 | hCoV-19/Netherlands/NB-RIVM-20398/2020\|EPI_ISL_728 | ···················· | ···················· | ···················· | ···················· | ···················· | ···················· |
| Y | 4893 | hCoV-19/Netherlands/NB-RIVM-20399/2020\|EPI_ISL_728 | ···················· | ···················· | ···················· | ···················· | ···················· | ···················· |
| Y | 4894 | hCoV-19/Netherlands/UT-RIVM-20400/2020\|EPI_ISL_728 | ···················· | ···················· | ···················· | ···················· | ···················· | ···················· |
| Y | 4895 | hCoV-19/Netherlands/UT-RIVM-20401/2020\|EPI_ISL_728 | ···················· | ···················· | ···················· | ···················· | ···················· | ···················· |
| Y | 4896 | hCoV-19/Netherlands/UT-RIVM-20402/2020\|EPI_ISL_728 | ···················· | ···················· | ···················· | ···················· | ···················· | ···················· |
| Y | 4897 | hCoV-19/Netherlands/UT-RIVM-20403/2020\|EPI_ISL_728 | ···················· | ···················· | ···················· | ···················· | ···················· | ···················· |
| Y | 4898 | hCoV-19/Netherlands/UT-RIVM-20404/2020\|EPI_ISL_728 | ···················· | ···················· | ···················· | ···················· | ···················· | ···················· |
| Y | 4899 | hCoV-19/Netherlands/UT-RIVM-20405/2020\|EPI_ISL_728 | ···················· | ···················· | ···················· | ···················· | ···················· | ···················· |
| Y | 4900 | hCoV-19/Netherlands/UT-RIVM-20406/2020\|EPI_ISL_728 | ···················· | ···················· | ···················· | ···················· | ······T············· | ···················· |
| Y | 4901 | hCoV-19/Netherlands/UT-RIVM-20407/2020\|EPI_ISL_728 | ···················· | ···················· | ···················· | ···················· | ···················· | ···················· |
| Y | 4902 | hCoV-19/Netherlands/DR-RIVM-20408/2020\|EPI_ISL_728 | ···················· | ···················· | ···················· | ···················· | ···················· | ···················· |
| Y | 4903 | hCoV-19/Netherlands/UT-RIVM-20409/2020\|EPI_ISL_728 | ···················· | ···················· | ···················· | ···················· | ···················· | ···················· |
| Y | 4904 | hCoV-19/Netherlands/NH-RIVM-20430/2020\|EPI_ISL_728 | ···················· | ···················· | ···················· | ···················· | ···················· | ···················· |
| Y | 4905 | hCoV-19/Netherlands/UT-RIVM-20431/2020\|EPI_ISL_728 | ···················· | ···················· | ···················· | ···················· | ···················· | ···················· |
| Y | 4906 | hCoV-19/Netherlands/NH-RIVM-20433/2020\|EPI_ISL_728 | ···················· | ···················· | ···················· | ···················· | ···················· | ···················· |
| Y | 4907 | hCoV-19/Netherlands/NH-RIVM-20434/2020\|EPI_ISL_728 | ···················· | ···················· | ···················· | ···················· | ···················· | ···················· |
| Y | 4908 | hCoV-19/Netherlands/NH-RIVM-20435/2020\|EPI_ISL_728 | ···················· | ···················· | ···················· | ···················· | ···················· | ···················· |
| Y | 4909 | hCoV-19/Netherlands/NH-RIVM-20436/2020\|EPI_ISL_728 | ···················· | ···················· | ···················· | ···················· | ···················· | ···················· |
| Y | 4910 | hCoV-19/Netherlands/NH-RIVM-20437/2020\|EPI_ISL_728 | ···················· | ···················· | ···················· | ···················· | ···················· | ···················· |
| Y | 4911 | hCoV-19/Netherlands/NH-RIVM-20438/2020\|EPI_ISL_728 | ···················· | ···················· | ···················· | ···················· | ···················· | ···················· |
| Y | 4912 | hCoV-19/Netherlands/NH-RIVM-20439/2020\|EPI_ISL_728 | ···················· | ···················· | ···················· | ···················· | ···················· | ···················· |
| Y | 4913 | hCoV-19/Netherlands/NH-RIVM-20440/2020\|EPI_ISL_728 | ···················· | ···················· | ···················· | ···················· | ···················· | ···················· |
| Y | 4914 | hCoV-19/Peru/LIM-UPCH-0242/2020\|EPI_ISL_729900\|202 | ···················· | ···················· | ···················· | ···················· | ···················· | ···················· |
| Y | 4915 | hCoV-19/Peru/LIM-UPCH-0243/2020\|EPI_ISL_729901\|202 | ···················· | ···················· | ···················· | ···················· | ···················· | ···················· |
| Y | 4916 | hCoV-19/Peru/AMZ-UPCH-0256/2020\|EPI_ISL_729902\|202 | ···················· | ···················· | ···················· | ···················· | ···················· | ···················· |
| Y | 4917 | hCoV-19/Peru/AMZ-UPCH-0257/2020\|EPI_ISL_729903\|202 | ···················· | ···················· | ···················· | ···················· | ···················· | ···················· |
| Y | 4918 | hCoV-19/Peru/AMZ-UPCH-0258/2020\|EPI_ISL_729904\|202 | ···················· | ····A·A············· | ···················· | ···················· | ···················· | ···················· |
| Y | 4919 | hCoV-19/Peru/AMZ-UPCH-0259/2020\|EPI_ISL_729905\|202 | ···················· | ···················· | ···················· | ···················· | ···················· | ···················· |
| Y | 4920 | hCoV-19/Peru/AMZ-UPCH-0260/2020\|EPI_ISL_729906\|202 | ···················· | ···················· | ···················· | ···················· | ···················· | ···················· |
| Y | 4921 | hCoV-19/Peru/AMZ-UPCH-0261/2020\|EPI_ISL_729907\|202 | ···················· | ···················· | ···················· | ···················· | ···················· | ···················· |
| Y | 4922 | hCoV-19/Peru/AMZ-UPCH-0262/2020\|EPI_ISL_729908\|202 | ···················· | ···················· | ···················· | ···················· | ···················· | ···················· |

| | | | | | | | | |
|---|---|---|---|---|---|---|---|---|
| Y | 4923 hCoV-19/Peru/AMZ-UPCH-0263/2020\|EPI_ISL_729909\|202 | ···················· | ···················· | ···················· | ···················· | ···················· | ···················· | |
| Y | 4924 hCoV-19/Peru/AMZ-UPCH-0264/2020\|EPI_ISL_729910\|202 | ···················· | ···················· | ···················· | ···················· | ···················· | ···················· | |
| Y | 4925 hCoV-19/Peru/AMZ-UPCH-0265/2020\|EPI_ISL_729911\|202 | ···················· | ···················· | ···················· | ···················· | ···················· | ···················· | |
| Y | 4926 hCoV-19/Peru/AMZ-UPCH-0266/2020\|EPI_ISL_729912\|202 | ···················· | ···················· | ···················· | ···················· | ···················· | ···················· | |
| Y | 4927 hCoV-19/Peru/AMZ-UPCH-0267/2020\|EPI_ISL_729913\|202 | ···················· | ···················· | ···················· | ···················· | ···················· | ···················· | |
| Y | 4928 hCoV-19/Peru/AMZ-UPCH-0268/2020\|EPI_ISL_729914\|202 | ···················· | ···················· | ···················· | ···················· | ···················· | ···················· | |
| Y | 4929 hCoV-19/Peru/AMZ-UPCH-0270/2020\|EPI_ISL_729915\|202 | ···················· | ···················· | ···················· | ···················· | ···················· | ···················· | |
| Y | 4930 hCoV-19/Peru/AMZ-UPCH-0272/2020\|EPI_ISL_729916\|202 | ···················· | ···················· | ···················· | ···················· | ···················· | ···················· | |
| Y | 4931 hCoV-19/Peru/AMZ-UPCH-0273/2020\|EPI_ISL_729917\|202 | ···················· | ···················· | ···················· | ···················· | ···················· | ···················· | |
| Y | 4932 hCoV-19/Peru/AMZ-UPCH-0274/2020\|EPI_ISL_729918\|202 | ···················· | ····A·A············· | ···················· | ···················· | ···················· | ···················· | |
| Y | 4933 hCoV-19/Peru/AMZ-UPCH-0275/2020\|EPI_ISL_729919\|202 | ···················· | ···················· | ···················· | ···················· | ···················· | ···················· | |
| Y | 4934 hCoV-19/Nigeria/KD-CV233/2020\|EPI_ISL_729920\|2020- | ···················· | ···················· | ···················· | ···················· | ···················· | ···················· | |
| Y | 4935 hCoV-19/Nigeria/NI-CV237/2020\|EPI_ISL_729923\|2020- | ···················· | ···················· | ···················· | ···················· | ···················· | ···················· | |
| Y | 4936 hCoV-19/Nigeria/FC-CV238/2020\|EPI_ISL_729924\|2020- | ···················· | ···················· | ···················· | ···················· | ···················· | ···················· | |
| Y | 4937 hCoV-19/Nigeria/NA-CV239/2020\|EPI_ISL_729925\|2020- | ···················· | ···················· | ···················· | ···················· | ···················· | ···················· | |
| Y | 4938 hCoV-19/Nigeria/NA-CV240/2020\|EPI_ISL_729926\|2020- | ···················· | ···················· | ···················· | ···················· | ···················· | ···················· | |
| Y | 4939 hCoV-19/Nigeria/BE-CV243/2020\|EPI_ISL_729928\|2020- | ···················· | ···················· | ···················· | ···················· | ···················· | ···················· | |
| Y | 4940 hCoV-19/Nigeria/NI-CV244/2020\|EPI_ISL_729929\|2020- | ···················· | ···················· | ···················· | ···················· | ···················· | ···················· | |
| Y | 4941 hCoV-19/Nigeria/FC-CV245/2020\|EPI_ISL_729930\|2020- | ···················· | ···················· | ···················· | ···················· | ···················· | ···················· | |
| Y | 4942 hCoV-19/Nigeria/FC-CV246/2020\|EPI_ISL_729931\|2020- | ···················· | ···················· | ···················· | ···················· | ···················· | ···················· | |
| Y | 4943 hCoV-19/Nigeria/FC-CV247/2020\|EPI_ISL_729932\|2020- | ···················· | ···················· | ···················· | ···················· | ···················· | ···················· | |
| Y | 4944 hCoV-19/Nigeria/FC-CV250/2020\|EPI_ISL_729935\|2020- | ···················· | ···················· | ···················· | ···················· | ···················· | ···················· | |
| Y | 4945 hCoV-19/Nigeria/KN-CV252/2020\|EPI_ISL_729937\|2020- | ···················· | ···················· | ···················· | ···················· | ···················· | ···················· | |
| Y | 4946 hCoV-19/Nigeria/LA-CV253/2020\|EPI_ISL_729938\|2020- | ···················· | ···················· | ···················· | ···················· | ···················· | ···················· | |
| Y | 4947 hCoV-19/Nigeria/LA-CV255/2020\|EPI_ISL_729939\|2020- | ···················· | ···················· | ···················· | ···················· | ···················· | ···················· | |
| Y | 4948 hCoV-19/Nigeria/LA-CV256/2020\|EPI_ISL_729940\|2020- | ···················· | ···················· | ···················· | ···················· | ···················· | ···················· | |
| Y | 4949 hCoV-19/Nigeria/LA-CV257/2020\|EPI_ISL_729941\|2020- | ···················· | ···················· | ···················· | ···················· | ···················· | ···················· | |
| Y | 4950 hCoV-19/Nigeria/KO-CV258/2020\|EPI_ISL_729942\|2020- | ···················· | ···················· | ···················· | ···················· | ···················· | ···················· | |
| Y | 4951 hCoV-19/Nigeria/LA-CV259/2020\|EPI_ISL_729943\|2020- | ···················· | ···················· | ···················· | ···················· | ···················· | ···················· | |
| Y | 4952 hCoV-19/Nigeria/OG-CV260/2020\|EPI_ISL_729944\|2020- | ···················· | ···················· | ···················· | ···················· | ···················· | ···················· | |
| Y | 4953 hCoV-19/Nigeria/LA-CV261/2020\|EPI_ISL_729945\|2020- | ···················· | ···················· | ···················· | ···················· | ···················· | ···················· | |
| Y | 4954 hCoV-19/Nigeria/LA-CV262/2020\|EPI_ISL_729946\|2020- | ···················· | ···················· | ···················· | ···················· | ···················· | ···················· | |
| Y | 4955 hCoV-19/Nigeria/LA-CV263/2020\|EPI_ISL_729947\|2020- | ···················· | ····A··············· | ···················· | ···················· | ···················· | ···················· | |
| Y | 4956 hCoV-19/Nigeria/LA-CV264/2020\|EPI_ISL_729948\|2020- | ···················· | ···················· | ···················· | ···················· | ···················· | ···················· | |
| Y | 4957 hCoV-19/Nigeria/LA-CV266/2020\|EPI_ISL_729950\|2020- | ···················· | ···················· | ···················· | ···················· | ···················· | ···················· | |
| Y | 4958 hCoV-19/Nigeria/AK-CV267/2020\|EPI_ISL_729951\|2020- | ···················· | ···················· | ···················· | ···················· | ···················· | ···················· | |
| Y | 4959 hCoV-19/Nigeria/AK-CV268/2020\|EPI_ISL_729952\|2020- | ···················· | ···················· | ···················· | ···················· | ···················· | ···················· | |
| Y | 4960 hCoV-19/Nigeria/AK-CV269/2020\|EPI_ISL_729953\|2020- | ···················· | ···················· | ···················· | ···················· | ···················· | ···················· | |
| Y | 4961 hCoV-19/Nigeria/AK-CV270/2020\|EPI_ISL_729954\|2020- | ···················· | ···················· | ···················· | ···················· | ···················· | ···················· | |
| Y | 4962 hCoV-19/Nigeria/AK-CV271/2020\|EPI_ISL_729955\|2020- | ···················· | ···················· | ···················· | ···················· | ···················· | ···················· | |
| Y | 4963 hCoV-19/Nigeria/AK-CV272/2020\|EPI_ISL_729956\|2020- | ···················· | ···················· | ···················· | ···················· | ···················· | ···················· | |
| Y | 4964 hCoV-19/Nigeria/AK-CV273/2020\|EPI_ISL_729957\|2020- | ···················· | ···················· | ···················· | ···················· | ···················· | ···················· | |
| Y | 4965 hCoV-19/Nigeria/BO-CV275/2020\|EPI_ISL_729959\|2020- | ···················· | ···················· | ···················· | ···················· | ···················· | ···················· | |
| Y | 4966 hCoV-19/Nigeria/AK-CV321/2020\|EPI_ISL_729992\|2020- | ···················· | ···················· | ···················· | ···················· | ···················· | ···················· | |
| Y | 4967 hCoV-19/Nigeria/AK-CV322/2020\|EPI_ISL_729993\|2020- | ···················· | ···················· | ···················· | ···················· | ···················· | ···················· | |
| Y | 4968 hCoV-19/England/ALDP-C398FB/2020\|EPI_ISL_730903\|20 | ···················· | ···················· | ···················· | ···················· | ···················· | ···················· | |
| Y | 4969 hCoV-19/England/ALDP-C3D657/2020\|EPI_ISL_730904\|20 | ···················· | ···················· | ···················· | ···················· | ···················· | ···················· | |
| Y | 4970 hCoV-19/England/ALDP-C39FC9/2020\|EPI_ISL_730906\|20 | ···················· | ···················· | ···················· | ···················· | ···················· | ···················· | |
| Y | 4971 hCoV-19/England/QEUH-C30DE6/2020\|EPI_ISL_730910\|20 | ···················· | ···················· | ···················· | ···················· | ···················· | ···················· | |
| Y | 4972 hCoV-19/England/QEUH-C30F77/2020\|EPI_ISL_730911\|20 | ···················· | ···················· | ···················· | ···················· | ···················· | ···················· | |
| Y | 4973 hCoV-19/England/ALDP-C39D65/2020\|EPI_ISL_730912\|20 | ···················· | ···················· | ···················· | ···················· | ···················· | ···················· | |
| Y | 4974 hCoV-19/England/QEUH-C32212/2020\|EPI_ISL_730913\|20 | ··················T· | ···················· | ···················· | ···················· | ···················· | ···················· | 1 |
| Y | 4975 hCoV-19/England/ALDP-C3DD8F/2020\|EPI_ISL_730914\|20 | ···················· | ···················· | ···················· | ···················· | ···················· | ···················· | |
| Y | 4976 hCoV-19/England/ALDP-C3B5A7/2020\|EPI_ISL_730915\|20 | ···················· | ···················· | ···················· | ···················· | ···················· | ···················· | |
| Y | 4977 hCoV-19/England/ALDP-C3A168/2020\|EPI_ISL_730916\|20 | ···················· | ···················· | ···················· | ···················· | ···················· | ···················· | |
| Y | 4978 hCoV-19/England/ALDP-C3C773/2020\|EPI_ISL_730917\|20 | ···················· | ···················· | ···················· | ···················· | ······T············· | ···················· | |
| Y | 4979 hCoV-19/England/ALDP-C3C755/2020\|EPI_ISL_730919\|20 | ···················· | ···················· | ···················· | ···················· | ···················· | ···················· | |
| Y | 4980 hCoV-19/England/ALDP-C3C913/2020\|EPI_ISL_730920\|20 | ···················· | ···················· | ···················· | ···················· | ···················· | ···················· | |
| Y | 4981 hCoV-19/England/ALDP-C3CF7B/2020\|EPI_ISL_730923\|20 | ···················· | ···················· | ···················· | ···················· | ···················· | ···················· | |
| Y | 4982 hCoV-19/England/ALDP-C3B6D1/2020\|EPI_ISL_730925\|20 | ···················· | ·A·················· | ···················· | ···················· | ···················· | ···A················ | |
| Y | 4983 hCoV-19/England/ALDP-C3B7CF/2020\|EPI_ISL_730927\|20 | ···················· | ···················· | ···················· | ···················· | ···················· | ···················· | |
| Y | 4984 hCoV-19/England/ALDP-C3D1FC/2020\|EPI_ISL_730928\|20 | ···················· | ···················· | ···················· | ···················· | ···················· | ···················· | |
| Y | 4985 hCoV-19/England/ALDP-C3CF8A/2020\|EPI_ISL_730930\|20 | ···················· | ···················· | ···················· | ···················· | ···················· | ···················· | |
| Y | 4986 hCoV-19/England/ALDP-C3B808/2020\|EPI_ISL_730931\|20 | ···················· | ···················· | ···················· | ···················· | ···················· | ···················· | |
| Y | 4987 hCoV-19/England/ALDP-C3D077/2020\|EPI_ISL_730933\|20 | ···················· | ···················· | ···················· | ···················· | ···················· | ···················· | |
| Y | 4988 hCoV-19/England/ALDP-C3D262/2020\|EPI_ISL_730934\|20 | ···················· | ···················· | ···················· | ···················· | ···················· | ···················· | |
| Y | 4989 hCoV-19/England/ALDP-C3D095/2020\|EPI_ISL_730940\|20 | ···················· | ···················· | ···················· | ···················· | ···················· | ···················· | |
| Y | 4990 hCoV-19/England/ALDP-C3B826/2020\|EPI_ISL_730974\|20 | ···················· | ···················· | ···················· | ···················· | ···················· | ···················· | |
| Y | 4991 hCoV-19/England/ALDP-C3D208/2020\|EPI_ISL_730975\|20 | ···················· | ···················· | ···················· | ···················· | ···················· | ···················· | |
| Y | 4992 hCoV-19/England/ALDP-C3B9F6/2020\|EPI_ISL_730976\|20 | ···················· | ···················· | ···················· | ···················· | ···················· | ···················· | |
| Y | 4993 hCoV-19/England/ALDP-C3C51F/2020\|EPI_ISL_730977\|20 | ···················· | ···················· | ···················· | ···················· | ······T············· | ···················· | |
| Y | 4994 hCoV-19/England/ALDP-C3C904/2020\|EPI_ISL_730978\|20 | ···················· | ···················· | ···················· | ···················· | ···················· | ···················· | |
| Y | 4995 hCoV-19/England/ALDP-C3C7A0/2020\|EPI_ISL_730979\|20 | ···················· | ···················· | ···················· | ···················· | ···················· | ···················· | |
| Y | 4996 hCoV-19/England/ALDP-C3A2ED/2020\|EPI_ISL_730980\|20 | ···················· | ···················· | ···················· | ···················· | ···················· | ···················· | |
| Y | 4997 hCoV-19/Russia/MOW-RII-MH3353S/2020\|EPI_ISL_733100 | ···················· | ···················· | ···················· | ···················· | ···················· | ···················· | |
| Y | 4998 hCoV-19/Russia/SE-RII-MH3355S/2020\|EPI_ISL_733102\| | ···················· | ···················· | ···················· | ···················· | ···················· | ···················· | |
| Y | 4999 hCoV-19/Russia/SE-RII-MH3356S/2020\|EPI_ISL_733103\| | ···················· | ···················· | ···················· | ···················· | ···················· | ···················· | |
| Y | 5000 hCoV-19/Russia/RYA-RII-MH3360S/2020\|EPI_ISL_733105 | ···················· | ···················· | ···················· | ···················· | ···················· | ···················· | |
| Y | 5001 hCoV-19/Russia/RYA-RII-MH3361S/2020\|EPI_ISL_733106 | ···················· | ···················· | ···················· | ···················· | ···················· | ···················· | |
| Y | 5002 hCoV-19/Russia/TUL-RII-MH3372S/2020\|EPI_ISL_733113 | ···················· | ···················· | ···················· | ···················· | ···················· | ···················· | |

Y 5003 hCoV-19/Russia/ROS-RII-MH3377S/2020|EPI_ISL_733117 ···················· ···················· ····················· ···················· ···················· ····················
Y 5004 hCoV-19/Russia/YAR-RII-MH3382S/2020|EPI_ISL_733121 ···················· ···················· ····················· ···················· ···················· ····················
Y 5005 hCoV-19/Russia/MOS-RII-MH3393S/2020|EPI_ISL_733126 ···················· ···················· ····················· ···················· ···················· ····················
Y 5006 hCoV-19/Russia/MOW-RII-MH3395S/2020|EPI_ISL_733128 ···················· ···················· ····················· ···················· ···················· ····················
Y 5007 hCoV-19/Russia/MOS-RII-MH3397S/2020|EPI_ISL_733129 ···················· ···················· ····················· ···················· ···················· ····················
Y 5008 hCoV-19/Russia/MOW-RII-MH3401S/2020|EPI_ISL_733132 ···················· ···················· ·········T··········· ···················· ···················· ····················
Y 5009 hCoV-19/Russia/STA-RII-MH3407S/2020|EPI_ISL_733135 ···················· ···················· ····················· ···················· ···················· ····················
Y 5010 hCoV-19/Russia/KO-RII-MH3409S/2020|EPI_ISL_733137| ···················· ···················· ····················· ···················· ···················· ····················
Y 5011 hCoV-19/Russia/KHM-RII-MH3413S/2020|EPI_ISL_733138 ···················· ···················· ····················· ···················· ···················· ····················
Y 5012 hCoV-19/Russia/KHM-RII-MH3415S/2020|EPI_ISL_733140 ···················· ···················· ····················· ···················· ···················· ····················
Y 5013 hCoV-19/Russia/KLU-RII-MH3421S/2020|EPI_ISL_733142 ···················· ···················· ····················· ···················· ···················· ····················
Y 5014 hCoV-19/Russia/KLU-RII-MH3423S/2020|EPI_ISL_733144 ···················· ···················· ····················· ···················· ···················· ····················
Y 5015 hCoV-19/Russia/MOS-RII-MH3424S/2020|EPI_ISL_733145 ···················· ···················· ····················· ···················· ···················· ····················
Y 5016 hCoV-19/Russia/ROS-RII-MH3425S/2020|EPI_ISL_733146 ···················· ···················· ····················· ···················· ···················· ····················
Y 5017 hCoV-19/Russia/ORE-RII-MH3429S/2020|EPI_ISL_733147 ···················· ···················· ····················· ···················· ···················· ····················
Y 5018 hCoV-19/Russia/MOW-RII-MH3430S/2020|EPI_ISL_733148 ···················· ···················· ····················· ···················· ···················· ····················
Y 5019 hCoV-19/Crimea/RII-MH3437S/2020|EPI_ISL_733151|202 ···················· ···················· ····················· ···················· ···················· ····················
Y 5020 hCoV-19/Russia/ORE-RII-MH3441S/2020|EPI_ISL_733153 ···················· ···················· ····················· ···················· ···················· ····················
Y 5021 hCoV-19/Russia/MO-RII-MH3571S/2020|EPI_ISL_733154| ···················· ···················· ····················· ···················· ···················· ····················
Y 5022 hCoV-19/Russia/MOW-RII-MH4070S/2020|EPI_ISL_733156 ···················· ···················· ····················· ···················· ···················· ····················
Y 5023 hCoV-19/Russia/MOW-RII-MH4074S/2020|EPI_ISL_733158 ···················· ···················· ····················· ···················· ···················· ····················
Y 5024 hCoV-19/Russia/MOW-RII-MH4077S/2020|EPI_ISL_733160 ···················· ···················· ····················· ···················· ···················· ····················
Y 5026 hCoV-19/Russia/AST-RII-MH4130S/2020|EPI_ISL_733167 ···················· ···················· ····················· ···················· ···················· ····················
Y 5028 hCoV-19/Russia/AST-RII-MH4136S/2020|EPI_ISL_733170 ···················· ···················· ····················· ···················· ···················· ····················
Y 5030 hCoV-19/Russia/AST-RII-MH4151S/2020|EPI_ISL_733175 ···················· ···················· ····················· ···················· ···················· ····················
Y 5031 hCoV-19/Russia/AST-RII-MH4152S/2020|EPI_ISL_733176 ···················· ···················· ····················· ···················· ···················· ····················
Y 5032 hCoV-19/Russia/AST-RII-MH4155S/2020|EPI_ISL_733178 ···················· ···················· ····················· ···················· ···················· ····················
Y 5033 hCoV-19/Russia/AST-RII-MH4156S/2020|EPI_ISL_733179 ···················· ···················· ····················· ···················· ···················· ····················
Y 5034 hCoV-19/Russia/AST-RII-MH4157S/2020|EPI_ISL_733180 ···················· ···················· ····················· ···················· ···················· ····················
Y 5035 hCoV-19/Russia/AST-RII-MH4159S/2020|EPI_ISL_733181 ···················· ···················· ····················· ···················· ···················· ····················
Y 5036 hCoV-19/Russia/AST-RII-MH4160S/2020|EPI_ISL_733182 ···················· ···················· ····················· ···················· ···················· ····················
Y 5037 hCoV-19/Russia/AST-RII-MH4442S/2020|EPI_ISL_733189 ···················· ···················· ····················· ···················· ···················· ····················
Y 5038 hCoV-19/Russia/AST-RII-MH4444S/2020|EPI_ISL_733190 ···················· ···················· ····················· ···················· ···················· ····················
Y 5043 hCoV-19/Russia/AST-RII-MH4456S/2020|EPI_ISL_733197 ···················· ···················· ····················· ···················· ···················· ····················
Y 5044 hCoV-19/Russia/AST-RII-MH4457S/2020|EPI_ISL_733198 ···················· ···················· ····················· ···················· ···················· ····················
Y 5046 hCoV-19/USA/NY-Wadsworth-269590-01/2020|EPI_ISL_73 ···················· ·A·················· ····················· ···················· ···················· ····················
Y 5047 hCoV-19/USA/NY-Wadsworth-269605-01/2020|EPI_ISL_73 ···················· ·A·················· ····················· ···················· ···················· ····················
Y 5048 hCoV-19/USA/NY-Wadsworth-269609-01/2020|EPI_ISL_73 ···················· ·A·················· ····················· ···················· ···················· ····················
Y 5049 hCoV-19/USA/NY-Wadsworth-269613-01/2020|EPI_ISL_73 ···················· ·A·················· ····················· ···················· ···················· ····················
Y 5050 hCoV-19/USA/NY-Wadsworth-269616-01/2020|EPI_ISL_73 ···················· ·A·················· ····················· ···················· ···················· ····················
Y 5051 hCoV-19/USA/NY-Wadsworth-269619-01/2020|EPI_ISL_73 ···················· ·A·················· ····················· ···················· ···················· ····················
Y 5052 hCoV-19/USA/NY-Wadsworth-269621-01/2020|EPI_ISL_73 ···················· ·A·················· ····················· ···················· ···················· ····················
Y 5053 hCoV-19/USA/NY-Wadsworth-269624-01/2020|EPI_ISL_73 ···················· ·A·················· ····················· ···················· ···················· ····················
Y 5054 hCoV-19/USA/NY-Wadsworth-269631-01/2020|EPI_ISL_73 ···················· ·A·················· ····················· ···················· ···················· ····················
Y 5055 hCoV-19/USA/NY-Wadsworth-269633-01/2020|EPI_ISL_73 ···················· ·A·················· ····················· ···················· ···················· ····················
Y 5056 hCoV-19/USA/NY-Wadsworth-269634-01/2020|EPI_ISL_73 ···················· ·A·················· ····················· ···················· ···················· ····················
Y 5057 hCoV-19/USA/NY-Wadsworth-269636-01/2020|EPI_ISL_73 ···················· ·A·················· ····················· ···················· ···················· ····················
Y 5058 hCoV-19/USA/NY-Wadsworth-269645-01/2020|EPI_ISL_73 ···················· ·A·················· ····················· ···················· ···················· ····················
Y 5059 hCoV-19/USA/NY-Wadsworth-269647-01/2020|EPI_ISL_73 ···················· ·A·················· ····················· ···················· ···················· ····················
Y 5060 hCoV-19/USA/NY-Wadsworth-269648-01/2020|EPI_ISL_73 ···················· ·A·················· ····················· ···················· ···················· ····················
Y 5061 hCoV-19/USA/NY-Wadsworth-269649-01/2020|EPI_ISL_73 ···················· ·A·················· ····················· ···················· ···················· ····················
Y 5062 hCoV-19/USA/NY-Wadsworth-269651-01/2020|EPI_ISL_73 ···················· ·A·················· ····················· ···················· ···················· ····················
Y 5063 hCoV-19/USA/NY-Wadsworth-269659-01/2020|EPI_ISL_73 ···················· ·A·················· ····················· ···················· ···················· ····················
Y 5064 hCoV-19/USA/NY-Wadsworth-271146-01/2020|EPI_ISL_73 ···················· ·A·················· ····················· ···················· ···················· ····················
Y 5065 hCoV-19/USA/NY-Wadsworth-281376-01/2020|EPI_ISL_73 ···················· ·A·················· ····················· ···················· ···················· ····················
Y 5066 hCoV-19/USA/NY-Wadsworth-281377-01/2020|EPI_ISL_73 ···················· ·A·················· ····················· ···················· ···················· ····················
Y 5067 hCoV-19/USA/NY-Wadsworth-281378-01/2020|EPI_ISL_73 ···················· ·A·················· ····················· ···················· ···················· ····················
Y 5068 hCoV-19/USA/NY-Wadsworth-281379-01/2020|EPI_ISL_73 ···················· ·A·················· ····················· ···················· ···················· ····················
Y 5069 hCoV-19/USA/NY-Wadsworth-281380-01/2020|EPI_ISL_73 ···················· ·A·················· ····················· ···················· ···················· ····················
Y 5070 hCoV-19/USA/NY-Wadsworth-281382-01/2020|EPI_ISL_73 ···················· ·A·················· ····················· ···················· ···················· ····················
Y 5071 hCoV-19/USA/NY-Wadsworth-281383-01/2020|EPI_ISL_73 ···················· ·A·················· ····················· ···················· ···················· ····················
Y 5072 hCoV-19/USA/NY-Wadsworth-281385-01/2020|EPI_ISL_73 ···················· ·A·················· ····················· ···················· ···················· ····················
Y 5073 hCoV-19/USA/NY-Wadsworth-281386-01/2020|EPI_ISL_73 ···················· ·A·················· ····················· ···················· ···················· ····················
Y 5074 hCoV-19/USA/NY-Wadsworth-281387-01/2020|EPI_ISL_73 ···················· ·A·················· ····················· ···················· ···················· ····················
Y 5075 hCoV-19/USA/NY-Wadsworth-281388-01/2020|EPI_ISL_73 ···················· ·A·················· ····················· ···················· ···················· ····················
Y 5076 hCoV-19/USA/NY-Wadsworth-281389-01/2020|EPI_ISL_73 ···················· ·A·················· ····················· ···················· ···················· ····················
Y 5077 hCoV-19/USA/NY-Wadsworth-281390-01/2020|EPI_ISL_73 ···················· ·A·················· ····················· ···················· ···················· ····················
Y 5078 hCoV-19/USA/NY-Wadsworth-281391-01/2020|EPI_ISL_73 ···················· ·A·················· ····················· ···················· ···················· ····················
Y 5079 hCoV-19/USA/NY-Wadsworth-281392-01/2020|EPI_ISL_73 ···················· ·A·················· ····················· ···················· ···················· ····················
Y 5080 hCoV-19/USA/NY-Wadsworth-281393-01/2020|EPI_ISL_73 ···················· ·A·················· ····················· ···················· ···················· ····················
Y 5081 hCoV-19/USA/NY-Wadsworth-281396-01/2020|EPI_ISL_73 ···················· ·A·················· ····················· ···················· ···················· ····················
Y 5082 hCoV-19/USA/NY-Wadsworth-281397-01/2020|EPI_ISL_73 ···················· ·A·················· ····················· ···················· ···················· ····················
Y 5083 hCoV-19/USA/NY-Wadsworth-281398-01/2020|EPI_ISL_73 ···················· ·A·················· ····················· ···················· ···················· ····················
Y 5084 hCoV-19/USA/NY-Wadsworth-281399-01/2020|EPI_ISL_73 ···················· ·A·················· ····················· ···················· ···················· ····················
Y 5085 hCoV-19/USA/NY-Wadsworth-281400-01/2020|EPI_ISL_73 ···················· ·A·················· ····················· ···················· ···················· ····················
Y 5086 hCoV-19/USA/NY-Wadsworth-281402-01/2020|EPI_ISL_73 ···················· ·A·················· ····················· ···················· ···················· ····················
Y 5087 hCoV-19/USA/NY-Wadsworth-281403-01/2020|EPI_ISL_73 ···················· ·A·················· ····················· ···················· ···················· ····················
Y 5088 hCoV-19/USA/NY-Wadsworth-281404-01/2020|EPI_ISL_73 ···················· ·A·················· ····················· ···················· ···················· ····················
Y 5089 hCoV-19/USA/NY-Wadsworth-281405-01/2020|EPI_ISL_73 ···················· ·A·················· ····················· ···················· ···················· ····················
Y 5090 hCoV-19/USA/NY-Wadsworth-281406-01/2020|EPI_ISL_73 ···················· ·A·················· ····················· ···················· ···················· ····················

| | | | | | | | |
|---|---|---|---|---|---|---|---|
| Y | 5091 hCoV-19/USA/NY-Wadsworth-281407-01/2020\|EPI_ISL_73 | ···················· | ·A·················· | ···················· | ···················· | ···················· | ···················· |
| Y | 5092 hCoV-19/USA/NY-Wadsworth-281408-01/2020\|EPI_ISL_73 | ···················· | ·A·················· | ···················· | ···················· | ···················· | ···················· |
| Y | 5093 hCoV-19/USA/NY-Wadsworth-281409-01/2020\|EPI_ISL_73 | ···················· | ·A·················· | ···················· | ···················· | ···················· | ···················· |
| Y | 5094 hCoV-19/USA/NY-Wadsworth-281411-01/2020\|EPI_ISL_73 | ···················· | ·A·················· | ···················· | ···················· | ···················· | ···················· |
| Y | 5095 hCoV-19/USA/NY-Wadsworth-281412-01/2020\|EPI_ISL_73 | ···················· | ·A·················· | ···················· | ···················· | ···················· | ···················· |
| Y | 5096 hCoV-19/USA/NY-Wadsworth-281413-01/2020\|EPI_ISL_73 | ···················· | ·A·················· | ···················· | ···················· | ···················· | ···················· |
| Y | 5097 hCoV-19/USA/NY-Wadsworth-281414-01/2020\|EPI_ISL_73 | ···················· | ·A·················· | ···················· | ···················· | ···················· | ···················· |
| Y | 5098 hCoV-19/USA/NY-Wadsworth-281415-01/2020\|EPI_ISL_73 | ···················· | ·A·················· | ···················· | ···················· | ···················· | ···················· |
| Y | 5099 hCoV-19/USA/NY-Wadsworth-281420-01/2020\|EPI_ISL_73 | ···················· | ·A·················· | ···················· | ···················· | ···················· | ···················· |
| Y | 5100 hCoV-19/Belgium/rega-0511661/2020\|EPI_ISL_734492\|2 | ···················· | ···················· | ···················· | ···················· | ···················· | ···················· |
| Y | 5101 hCoV-19/Belgium/rega-0611919/2020\|EPI_ISL_734493\|2 | ···················· | ···················· | ···················· | ···················· | ···················· | ···················· |
| Y | 5102 hCoV-19/Belgium/rega-0714704/2020\|EPI_ISL_734494\|2 | ···················· | ···················· | ···················· | ···················· | ···················· | ···················· |
| Y | 5103 hCoV-19/Belgium/rega-10151117/2020\|EPI_ISL_734495\| | ···················· | ···················· | ···················· | ···················· | ···················· | ···················· |
| Y | 5104 hCoV-19/Belgium/rega-10191096/2020\|EPI_ISL_734496\| | ···················· | ···················· | ···················· | ···················· | ···················· | ···················· |
| Y | 5105 hCoV-19/Belgium/rega-10191273/2020\|EPI_ISL_734498\| | ···················· | ···················· | ···················· | ···················· | ···················· | ···················· |
| Y | 5106 hCoV-19/Belgium/rega-10191281/2020\|EPI_ISL_734499\| | ···················· | ···················· | ···················· | ···················· | ···················· | ···················· |
| Y | 5107 hCoV-19/England/MILK-C63B4A/2020\|EPI_ISL_735715\|20 | ···················· | ···················· | ···················· | ···················· | ···················· | ···················· |
| Y | 5108 hCoV-19/England/MILK-C61F7D/2020\|EPI_ISL_735718\|20 | ···················· | ···················· | ···················· | ···················· | ···················· | ···················· |
| Y | 5109 hCoV-19/England/MILK-C63A5C/2020\|EPI_ISL_735721\|20 | ···················· | ···················· | ···················· | ···················· | ···················· | ···················· |
| Y | 5110 hCoV-19/England/MILK-C6210D/2020\|EPI_ISL_735722\|20 | ···················· | ···················· | ···················· | ···················· | ······T············· | ···················· |
| Y | 5111 hCoV-19/England/MILK-C621FE/2020\|EPI_ISL_735725\|20 | ···················· | ···················· | ···················· | ···················· | ···················· | ···················· |
| Y | 5112 hCoV-19/England/MILK-C61EAD/2020\|EPI_ISL_735732\|20 | ···················· | ···················· | ···················· | ···················· | ···················· | ···················· |
| Y | 5113 hCoV-19/England/MILK-C61FB9/2020\|EPI_ISL_735764\|20 | ···················· | ···················· | ···················· | ···················· | ···················· | ···················· |
| Y | 5114 hCoV-19/England/MILK-C6165A/2020\|EPI_ISL_735769\|20 | ···················· | ···················· | ···················· | ···················· | ···················· | ···················· |
| Y | 5115 hCoV-19/England/MILK-C6111D/2020\|EPI_ISL_735777\|20 | ···················· | ···················· | ···················· | ···················· | ···················· | ···················· |
| Y | 5116 hCoV-19/England/MILK-C62219/2020\|EPI_ISL_735781\|20 | ···················· | ·A·················· | ···················· | ···················· | ···················· | ···················· |
| Y | 5117 hCoV-19/England/MILK-C615B7/2020\|EPI_ISL_735790\|20 | ···················· | ·A·················· | ···················· | ···················· | ···················· | ···A················ |
| Y | 5118 hCoV-19/England/MILK-C61247/2020\|EPI_ISL_735791\|20 | ···················· | ·A·················· | ···················· | ···················· | ···················· | ···A················ |
| Y | 5119 hCoV-19/England/MILK-C61881/2020\|EPI_ISL_735793\|20 | ···················· | ···················· | ···················· | ···················· | ······T············· | ···················· |
| Y | 5120 hCoV-19/England/MILK-C61D0A/2020\|EPI_ISL_735799\|20 | ···················· | ···················· | ···················· | ···················· | ···················· | ···················· |
| Y | 5121 hCoV-19/Wales/PHWC-493C28/2020\|EPI_ISL_726000\|2020 | ···················· | ···················· | ···················· | ···················· | ···················· | ···················· |
| Y | 5122 hCoV-19/Wales/PHWC-493C55/2020\|EPI_ISL_726003\|2020 | ···················· | ···················· | ···················· | ···················· | ···················· | ···················· |
| Y | 5123 hCoV-19/Wales/PHWC-493C64/2020\|EPI_ISL_726004\|2020 | ···················· | ···················· | ···················· | ···················· | ···················· | ···················· |
| Y | 5124 hCoV-19/Wales/PHWC-493C73/2020\|EPI_ISL_726005\|2020 | ···················· | ···················· | ···················· | ···················· | ···················· | ···················· |
| Y | 5125 hCoV-19/Wales/PHWC-493C82/2020\|EPI_ISL_726006\|2020 | ···················· | ···················· | ···················· | ···················· | ···················· | ···················· |
| Y | 5127 hCoV-19/Wales/PHWC-493CDD/2020\|EPI_ISL_726011\|2020 | ···················· | ···················· | ···················· | ···················· | ···················· | ···················· |
| Y | 5128 hCoV-19/Wales/PHWC-493CEC/2020\|EPI_ISL_726012\|2020 | ···················· | ···················· | ···················· | ···················· | ···················· | ···················· |
| Y | 5129 hCoV-19/Wales/PHWC-493CFB/2020\|EPI_ISL_726013\|2020 | ···················· | ···················· | ···················· | ···················· | ···················· | ···················· |
| Y | 5130 hCoV-19/Wales/PHWC-493D07/2020\|EPI_ISL_726014\|2020 | ···················· | ···················· | ···················· | ···················· | ···················· | ···················· |
| Y | 5131 hCoV-19/Wales/PHWC-493D25/2020\|EPI_ISL_726016\|2020 | ···················· | ···················· | ···················· | ···················· | ···················· | ···················· |
| Y | 5132 hCoV-19/Wales/PHWC-493D34/2020\|EPI_ISL_726017\|2020 | ···················· | ···················· | ···················· | ···················· | ···················· | ···················· |
| Y | 5133 hCoV-19/Wales/PHWC-493D43/2020\|EPI_ISL_726018\|2020 | ···················· | ···················· | ···················· | ···················· | ···················· | ···················· |
| Y | 5134 hCoV-19/Wales/PHWC-493D52/2020\|EPI_ISL_726019\|2020 | ···················· | ···················· | ···················· | ···················· | ···················· | ···················· |
| Y | 5135 hCoV-19/Wales/PHWC-493D61/2020\|EPI_ISL_726020\|2020 | ···················· | ···················· | ···················· | ···················· | ···················· | ···················· |
| Y | 5136 hCoV-19/Wales/PHWC-493D70/2020\|EPI_ISL_726021\|2020 | ···················· | ···················· | ···················· | ···················· | ···················· | ···················· |
| Y | 5137 hCoV-19/Wales/PHWC-493D8F/2020\|EPI_ISL_726022\|2020 | ···················· | ···················· | ···················· | ···················· | ····T··············· | ···················· |
| Y | 5138 hCoV-19/Wales/PHWC-493D9E/2020\|EPI_ISL_726023\|2020 | ···················· | ···················· | ···················· | ···················· | ···················· | ···················· |
| Y | 5141 hCoV-19/Wales/PHWC-493DE9/2020\|EPI_ISL_726028\|2020 | ···················· | ···················· | ···················· | ···················· | ···················· | ···················· |
| Y | 5142 hCoV-19/Wales/PHWC-493DF8/2020\|EPI_ISL_726029\|2020 | ···················· | ···················· | ···················· | ···················· | ···················· | ···················· |
| Y | 5144 hCoV-19/Wales/PHWC-493E13/2020\|EPI_ISL_726031\|2020 | ···················· | ···················· | ···················· | ···················· | ···················· | ···················· |
| Y | 5145 hCoV-19/Wales/PHWC-493E5F/2020\|EPI_ISL_726033\|2020 | ···················· | ···················· | ···················· | ···················· | ···················· | ···················· |
| Y | 5146 hCoV-19/Wales/PHWC-493E6E/2020\|EPI_ISL_726034\|2020 | ···················· | ···················· | ···················· | ···················· | ···················· | ···················· |
| Y | 5148 hCoV-19/Wales/PHWC-493EF5/2020\|EPI_ISL_726038\|2020 | ···················· | ···················· | ···················· | ···················· | ···················· | ···················· |
| Y | 5149 hCoV-19/Wales/PHWC-493F01/2020\|EPI_ISL_726039\|2020 | ···················· | ···················· | ···················· | ···················· | ···················· | ···················· |
| Y | 5150 hCoV-19/Wales/PHWC-493F10/2020\|EPI_ISL_726040\|2020 | ···················· | ···················· | ···················· | ···················· | ···················· | ···················· |
| Y | 5151 hCoV-19/Wales/PHWC-493F2F/2020\|EPI_ISL_726041\|2020 | ···················· | ···················· | ···················· | ···················· | ···················· | ···················· |
| Y | 5152 hCoV-19/Wales/PHWC-493F3E/2020\|EPI_ISL_726042\|2020 | ···················· | ···················· | ···················· | ···················· | ···················· | ···················· |
| Y | 5153 hCoV-19/Wales/PHWC-493F4D/2020\|EPI_ISL_726043\|2020 | ···················· | ···················· | ···················· | ···················· | ···················· | ···················· |
| Y | 5154 hCoV-19/Wales/PHWC-493F5C/2020\|EPI_ISL_726044\|2020 | ···················· | ···················· | ···················· | ···················· | ···················· | ···················· |
| Y | 5156 hCoV-19/Wales/PHWC-493F7A/2020\|EPI_ISL_726046\|2020 | ···················· | ···················· | ···················· | ···················· | ···················· | ···················· |
| Y | 5157 hCoV-19/Wales/PHWC-494155/2020\|EPI_ISL_726074\|2020 | ···················· | ···················· | ···················· | ···················· | ···················· | ···················· |
| Y | 5158 hCoV-19/Wales/PHWC-494164/2020\|EPI_ISL_726075\|2020 | ···················· | ···················· | ···················· | ···················· | ···················· | ···················· |
| Y | 5159 hCoV-19/Wales/PHWC-494173/2020\|EPI_ISL_726076\|2020 | ···················· | ···················· | ···················· | ···················· | ···················· | ···················· |
| Y | 5160 hCoV-19/Wales/PHWC-494182/2020\|EPI_ISL_726077\|2020 | ···················· | ···················· | ···················· | ···················· | ···················· | ···················· |
| Y | 5163 hCoV-19/Wales/PHWC-4993E1/2020\|EPI_ISL_727300\|2020 | ···················· | ···················· | ···················· | ···················· | ···················· | ···················· |
| Y | 5164 hCoV-19/Wales/PHWC-4993F0/2020\|EPI_ISL_727301\|2020 | ···················· | ···················· | ···················· | ···················· | ···················· | ···················· |
| Y | 5165 hCoV-19/Wales/PHWC-49940C/2020\|EPI_ISL_727302\|2020 | ···················· | ···················· | ···················· | ···················· | ···················· | ···················· |
| Y | 5166 hCoV-19/Wales/PHWC-49941B/2020\|EPI_ISL_727303\|2020 | ···················· | ···················· | ···················· | ···················· | ···················· | ···················· |
| Y | 5168 hCoV-19/Wales/PHWC-499457/2020\|EPI_ISL_727306\|2020 | ···················· | ···················· | ···················· | ···················· | ···················· | ···················· |
| Y | 5169 hCoV-19/Wales/PHWC-499466/2020\|EPI_ISL_727307\|2020 | ···················· | ···················· | ···················· | ···················· | ···················· | ···················· |
| Y | 5170 hCoV-19/Wales/PHWC-499475/2020\|EPI_ISL_727308\|2020 | ···················· | ···················· | ···················· | ···················· | ·NN················· | ···················· |
| Y | 5171 hCoV-19/Wales/PHWC-499484/2020\|EPI_ISL_727309\|2020 | ···················· | ···················· | ···················· | ···················· | ···················· | ···················· |
| Y | 5172 hCoV-19/Wales/PHWC-499493/2020\|EPI_ISL_727310\|2020 | ···················· | ···················· | ···················· | ···················· | ···················· | ···················· |
| Y | 5173 hCoV-19/Wales/PHWC-4994A2/2020\|EPI_ISL_727311\|2020 | ···················· | ···················· | ···················· | ···················· | ···················· | ···················· |
| Y | 5174 hCoV-19/Wales/PHWC-4994B1/2020\|EPI_ISL_727312\|2020 | ···················· | ···················· | ···················· | ···················· | ···················· | ···················· |
| Y | 5176 hCoV-19/Wales/PHWC-4994DF/2020\|EPI_ISL_727314\|2020 | ···················· | ···················· | ···················· | ···················· | ···················· | ···················· |
| Y | 5178 hCoV-19/Wales/PHWC-499509/2020\|EPI_ISL_727316\|2020 | ···················· | ···················· | ···················· | ···················· | ···················· | ···················· |
| Y | 5179 hCoV-19/Wales/PHWC-499518/2020\|EPI_ISL_727317\|2020 | ···················· | ···················· | ···················· | ···················· | ···················· | ···················· |
| Y | 5180 hCoV-19/Wales/PHWC-499536/2020\|EPI_ISL_727319\|2020 | ···················· | ···················· | ···················· | ···················· | ···················· | ···················· |
| Y | 5181 hCoV-19/Wales/PHWC-499606/2020\|EPI_ISL_727331\|2020 | ···················· | ···················· | ···················· | ···················· | ···················· | ···················· |

| | | | | | | | | | |
|---|---|---|---|---|---|---|---|---|---|
| Y | 5182 | hCoV-19/Wales/PHWC-499624/2020\|EPI_ISL_727333\|2020 | ···················· | ···················· | ···················· | ···················· | ···················· | ···················· | |
| Y | 5183 | hCoV-19/Wales/PHWC-499633/2020\|EPI_ISL_727334\|2020 | ···················· | ···················· | ···················· | ···················· | ···················· | ···················· | |
| Y | 5184 | hCoV-19/Wales/PHWC-499642/2020\|EPI_ISL_727335\|2020 | ···················· | ···················· | ···················· | ···················· | ···················· | ···················· | |
| Y | 5185 | hCoV-19/Wales/PHWC-499651/2020\|EPI_ISL_727336\|2020 | ···················· | ··········A········· | ···················· | ···················· | ···················· | ···················· | |
| Y | 5186 | hCoV-19/Wales/PHWC-49967F/2020\|EPI_ISL_727338\|2020 | ···················· | ···················· | ···················· | ···················· | ···················· | ···················· | |
| Y | 5187 | hCoV-19/Wales/PHWC-49968E/2020\|EPI_ISL_727339\|2020 | ···················· | ···················· | ···················· | ···················· | ···················· | ···················· | |
| Y | 5188 | hCoV-19/Wales/PHWC-4996AC/2020\|EPI_ISL_727340\|2020 | ···················· | ···················· | ···················· | ···················· | ···················· | ···················· | |
| Y | 5190 | hCoV-19/Wales/PHWC-4996D9/2020\|EPI_ISL_727343\|2020 | ···················· | ···················· | ···················· | ···················· | ···················· | ···················· | |
| Y | 5192 | hCoV-19/Wales/PHWC-499703/2020\|EPI_ISL_727345\|2020 | ···················· | ···················· | ···················· | ···················· | ···················· | ···················· | |
| Y | 5194 | hCoV-19/Wales/PHWC-499721/2020\|EPI_ISL_727347\|2020 | ···················· | ···················· | ···················· | ···················· | ···················· | ···················· | |
| Y | 5195 | hCoV-19/Wales/PHWC-49974F/2020\|EPI_ISL_727349\|2020 | ···················· | ···················· | ···················· | ···················· | ···················· | ···················· | |
| Y | 5196 | hCoV-19/Wales/PHWC-49975E/2020\|EPI_ISL_727350\|2020 | ···················· | ···················· | ···················· | ···················· | ···················· | ···················· | |
| Y | 5197 | hCoV-19/Wales/PHWC-49976D/2020\|EPI_ISL_727351\|2020 | ···················· | ···················· | ···················· | ···················· | ···················· | ···················· | |
| Y | 5198 | hCoV-19/Wales/PHWC-49977C/2020\|EPI_ISL_727352\|2020 | ···················· | ···················· | ···················· | ···················· | ···················· | ···················· | |
| Y | 5199 | hCoV-19/Wales/PHWC-49978B/2020\|EPI_ISL_727353\|2020 | ···················· | ···················· | ···················· | ···················· | ···················· | ···················· | |
| Y | 5201 | hCoV-19/Wales/PHWC-4997B8/2020\|EPI_ISL_727356\|2020 | ···················· | ···················· | ···················· | ···················· | ···················· | ···················· | |
| Y | 5202 | hCoV-19/Wales/PHWC-4997C7/2020\|EPI_ISL_727357\|2020 | ···················· | ···················· | ···················· | ···················· | ···················· | ···················· | |
| Y | 5204 | hCoV-19/Wales/PHWC-4997E5/2020\|EPI_ISL_727359\|2020 | ···················· | ···················· | ···················· | ···················· | ···················· | ···················· | |
| Y | 5205 | hCoV-19/Wales/PHWC-4997F4/2020\|EPI_ISL_727360\|2020 | ···················· | ···················· | ···················· | ···················· | NNN················· | N··················· | 1 |
| Y | 5206 | hCoV-19/Wales/PHWC-499800/2020\|EPI_ISL_727361\|2020 | ···················· | ···················· | ···················· | ···················· | ···················· | ···················· | |
| Y | 5208 | hCoV-19/Wales/PHWC-49982E/2020\|EPI_ISL_727363\|2020 | ···················· | ···················· | ···················· | ···················· | ···················· | ···················· | |
| Y | 5210 | hCoV-19/Wales/PHWC-49984C/2020\|EPI_ISL_727365\|2020 | ···················· | ···················· | ···················· | ···················· | ···················· | ···················· | |
| Y | 5211 | hCoV-19/Wales/PHWC-49985B/2020\|EPI_ISL_727366\|2020 | ···················· | ···················· | ···················· | ···················· | ···················· | ···················· | |
| Y | 5212 | hCoV-19/Wales/PHWC-49986A/2020\|EPI_ISL_727367\|2020 | ···················· | ···················· | ···················· | ···················· | ···················· | ···················· | |
| Y | 5214 | hCoV-19/Wales/PHWC-499888/2020\|EPI_ISL_727369\|2020 | ···················· | ···················· | ···················· | ···················· | ···················· | ···················· | |
| Y | 5218 | hCoV-19/Wales/PHWC-4998E2/2020\|EPI_ISL_727374\|2020 | ···················· | ···················· | ···················· | ···················· | ···················· | ···················· | |
| Y | 5219 | hCoV-19/Wales/PHWC-49990D/2020\|EPI_ISL_727376\|2020 | ···················· | ···················· | ···················· | ···················· | ···················· | ···················· | |
| Y | 5221 | hCoV-19/Wales/PHWC-49993A/2020\|EPI_ISL_727379\|2020 | ···················· | ···················· | ···················· | ···················· | ···················· | ···················· | |
| Y | 5222 | hCoV-19/Netherlands/ZH-RIVM-20410/2020\|EPI_ISL_728 | ···················· | ···················· | ···················· | ···················· | ······T············· | ···················· | |
| Y | 5223 | hCoV-19/Netherlands/UT-RIVM-20411/2020\|EPI_ISL_728 | ···················· | ···················· | ···················· | ···················· | ···················· | ···················· | |
| Y | 5224 | hCoV-19/Netherlands/ZH-RIVM-20412/2020\|EPI_ISL_728 | ···················· | ···················· | ···················· | ···················· | ···················· | ···················· | |
| Y | 5225 | hCoV-19/Netherlands/ZH-RIVM-20413/2020\|EPI_ISL_728 | ···················· | ···················· | ···················· | ···················· | ···················· | ···················· | |
| Y | 5226 | hCoV-19/Netherlands/ZH-RIVM-20414/2020\|EPI_ISL_728 | ···················· | ···················· | ···················· | ···················· | ···················· | ···················· | |
| Y | 5227 | hCoV-19/Netherlands/NB-RIVM-20415/2020\|EPI_ISL_728 | ···················· | ···················· | ···················· | ···················· | ···················· | ···················· | |
| Y | 5228 | hCoV-19/Netherlands/LI-RIVM-20416/2020\|EPI_ISL_728 | ···················· | ···················· | ···················· | ···················· | ···················· | ···················· | |
| Y | 5229 | hCoV-19/Netherlands/LI-RIVM-20417/2020\|EPI_ISL_728 | ···················· | ···················· | ···················· | ···················· | ···················· | ···················· | |
| Y | 5230 | hCoV-19/Netherlands/GR-RIVM-20418/2020\|EPI_ISL_728 | ···················· | ···················· | ···················· | ···················· | ···················· | ···················· | |
| Y | 5231 | hCoV-19/Netherlands/DR-RIVM-20419/2020\|EPI_ISL_728 | ···················· | ···················· | ···················· | ···················· | ···················· | ···················· | |
| Y | 5232 | hCoV-19/Netherlands/DR-RIVM-20420/2020\|EPI_ISL_728 | ···················· | ···················· | ···················· | ···················· | ···················· | ···················· | |
| Y | 5233 | hCoV-19/Netherlands/GR-RIVM-20421/2020\|EPI_ISL_728 | ···················· | ···················· | ···················· | ···················· | ···················· | ···················· | |
| Y | 5234 | hCoV-19/Netherlands/DR-RIVM-20422/2020\|EPI_ISL_728 | ···················· | ···················· | ···················· | ···················· | ······T············· | ···················· | |
| Y | 5235 | hCoV-19/Netherlands/DR-RIVM-20423/2020\|EPI_ISL_728 | ···················· | ···················· | ···················· | ···················· | ···················· | ···················· | |
| Y | 5236 | hCoV-19/Netherlands/GR-RIVM-20424/2020\|EPI_ISL_728 | ···················· | ···················· | ···················· | ···················· | ···················· | ···················· | |
| Y | 5237 | hCoV-19/Netherlands/GR-RIVM-20425/2020\|EPI_ISL_728 | ···················· | ···················· | ···················· | ···················· | ···················· | ···················· | |
| Y | 5238 | hCoV-19/Netherlands/FR-RIVM-20426/2020\|EPI_ISL_728 | ···················· | ···················· | ···················· | ···················· | ···················· | ···················· | |
| Y | 5239 | hCoV-19/Netherlands/GR-RIVM-20427/2020\|EPI_ISL_728 | ···················· | ···················· | ···················· | ···················· | ···················· | ···················· | |
| Y | 5240 | hCoV-19/Netherlands/GR-RIVM-20428/2020\|EPI_ISL_728 | ···················· | ···················· | ···················· | ···················· | ···················· | ···················· | |
| Y | 5241 | hCoV-19/Netherlands/NH-RIVM-20429/2020\|EPI_ISL_728 | ···················· | ···················· | ···················· | ···················· | ···················· | ···················· | |
| Y | 5242 | hCoV-19/Netherlands/ZH-RIVM-20441/2020\|EPI_ISL_728 | ···················· | ···················· | ···················· | ···················· | ······T············· | ···················· | |
| Y | 5243 | hCoV-19/Netherlands/ZH-RIVM-20442/2020\|EPI_ISL_728 | ···················· | ···················· | ···················· | ···················· | ······T············· | ···················· | |
| Y | 5244 | hCoV-19/Netherlands/LI-RIVM-20443/2020\|EPI_ISL_728 | ···················· | ···················· | ···················· | ···················· | ···················· | ···················· | |
| Y | 5245 | hCoV-19/Netherlands/LI-RIVM-20444/2020\|EPI_ISL_728 | ···················· | ···················· | ···················· | ···················· | ···················· | ···················· | |
| Y | 5246 | hCoV-19/Netherlands/LI-RIVM-20445/2020\|EPI_ISL_728 | ···················· | ···················· | ···················· | ···················· | ···················· | ···················· | |
| Y | 5247 | hCoV-19/Netherlands/LI-RIVM-20446/2020\|EPI_ISL_728 | ···················· | ···················· | ···················· | ···················· | ···················· | ···················· | |
| Y | 5248 | hCoV-19/Netherlands/LI-RIVM-20447/2020\|EPI_ISL_728 | ···················· | ···················· | ···················· | ···················· | ···················· | ···················· | |
| Y | 5249 | hCoV-19/Netherlands/LI-RIVM-20448/2020\|EPI_ISL_728 | ···················· | ···················· | ···················· | ···················· | ···················· | ···················· | |
| Y | 5250 | hCoV-19/Netherlands/ZH-RIVM-20449/2020\|EPI_ISL_728 | ···················· | ···················· | ···················· | ···················· | ···················· | ···················· | |
| Y | 5251 | hCoV-19/Netherlands/ZH-RIVM-20450/2020\|EPI_ISL_728 | ···················· | ···················· | ···················· | ···················· | ······T············· | ···················· | |
| Y | 5252 | hCoV-19/Netherlands/ZH-RIVM-20451/2020\|EPI_ISL_728 | ···················· | ···················· | ···················· | ···················· | ···················· | ···················· | |
| Y | 5253 | hCoV-19/Netherlands/ZH-RIVM-20452/2020\|EPI_ISL_728 | ···················· | ···················· | ···················· | ···················· | ···················· | ···················· | |
| Y | 5254 | hCoV-19/Netherlands/ZH-RIVM-20453/2020\|EPI_ISL_728 | ···················· | ···················· | ···················· | ···················· | ···················· | ···················· | |
| Y | 5255 | hCoV-19/Netherlands/ZH-RIVM-20454/2020\|EPI_ISL_728 | ···················· | ···················· | ···················· | ···················· | ···················· | ···················· | |
| Y | 5256 | hCoV-19/Netherlands/OV-RIVM-20458/2020\|EPI_ISL_728 | ···················· | ···················· | ···················· | ···················· | ···················· | ···················· | |
| Y | 5257 | hCoV-19/Netherlands/OV-RIVM-20459/2020\|EPI_ISL_728 | ···················· | ···················· | ···················· | ···················· | ···················· | ···················· | |
| Y | 5258 | hCoV-19/Netherlands/OV-RIVM-20461/2020\|EPI_ISL_728 | ···················· | ···················· | ···················· | ···················· | ···················· | ···················· | |
| Y | 5259 | hCoV-19/Netherlands/OV-RIVM-20462/2020\|EPI_ISL_728 | ···················· | ···················· | ···················· | ···················· | ···················· | ···················· | |
| Y | 5260 | hCoV-19/Netherlands/OV-RIVM-20463/2020\|EPI_ISL_728 | ···················· | ···················· | ···················· | ···················· | ···················· | ···················· | |
| Y | 5261 | hCoV-19/Netherlands/OV-RIVM-20464/2020\|EPI_ISL_728 | ···················· | ···················· | ···················· | ···················· | ···················· | ···················· | |
| Y | 5262 | hCoV-19/Netherlands/OV-RIVM-20465/2020\|EPI_ISL_728 | ···················· | ···················· | ···················· | ···················· | ···················· | ···················· | |
| Y | 5263 | hCoV-19/Netherlands/OV-RIVM-20466/2020\|EPI_ISL_728 | ···················· | ···················· | ···················· | ···················· | ···················· | ···················· | |
| Y | 5264 | hCoV-19/Netherlands/OV-RIVM-20467/2020\|EPI_ISL_728 | ···················· | ···················· | ···················· | ···················· | ···················· | ···················· | |
| Y | 5265 | hCoV-19/Netherlands/OV-RIVM-20468/2020\|EPI_ISL_728 | ···················· | ···················· | ···················· | ···················· | ···················· | ···················· | |
| Y | 5266 | hCoV-19/Netherlands/OV-RIVM-20469/2020\|EPI_ISL_728 | ···················· | ···················· | ···················· | ···················· | ···················· | ···················· | |
| Y | 5267 | hCoV-19/Netherlands/OV-RIVM-20470/2020\|EPI_ISL_728 | ···················· | ···················· | ···················· | ···················· | ···················· | ···················· | |
| Y | 5268 | hCoV-19/Netherlands/OV-RIVM-20471/2020\|EPI_ISL_728 | ···················· | ···················· | ···················· | ···················· | ···················· | ···················· | |
| Y | 5269 | hCoV-19/Netherlands/OV-RIVM-20472/2020\|EPI_ISL_728 | ···················· | ···················· | ···················· | ···················· | ···················· | ···················· | |
| Y | 5270 | hCoV-19/Netherlands/GE-RIVM-20473/2020\|EPI_ISL_728 | ···················· | ···················· | ···················· | ···················· | ···················· | ···················· | |
| Y | 5271 | hCoV-19/Netherlands/GE-RIVM-20474/2020\|EPI_ISL_728 | ···················· | ···················· | ···················· | ···················· | ···················· | ···················· | |
| Y | 5272 | hCoV-19/Netherlands/GE-RIVM-20475/2020\|EPI_ISL_728 | ···················· | ···················· | ···················· | ···················· | ···················· | ···················· | |
| Y | 5273 | hCoV-19/Netherlands/NB-RIVM-20476/2020\|EPI_ISL_728 | ···················· | ···················· | ···················· | ···················· | ···················· | ···················· | |

Y 5274 hCoV-19/Netherlands/NB-RIVM-20477/2020|EPI_ISL_728 ···················· ···················· ····················· ···················· ···················· ····················
Y 5275 hCoV-19/Netherlands/LI-RIVM-20478/2020|EPI_ISL_728 ···················· ···················· ····················· ···················· ···················· ····················
Y 5276 hCoV-19/Netherlands/LI-RIVM-20479/2020|EPI_ISL_728 ···················· ···················· ····················· ···················· ···················· ····················
Y 5277 hCoV-19/Netherlands/GR-RIVM-20480/2020|EPI_ISL_728 ···················· ···················· ····················· ···················· ···················· ····················
Y 5278 hCoV-19/Netherlands/GR-RIVM-20481/2020|EPI_ISL_728 ···················· ···················· ····················· ···················· ···················· ····················
Y 5279 hCoV-19/Netherlands/NH-RIVM-20482/2020|EPI_ISL_728 ···················· ···················· ····················· ···················· ···················· ····················
Y 5280 hCoV-19/Netherlands/NH-RIVM-20483/2020|EPI_ISL_728 ···················· ···················· ····················· ···················· ······T············· ····················
Y 5281 hCoV-19/Netherlands/NH-RIVM-20484/2020|EPI_ISL_728 ···················· ···················· ····················· ···················· ········C··········· ····················
Y 5282 hCoV-19/Nigeria/BO-CV276/2020|EPI_ISL_729960|2020- ···················· ···················· ····················· ···················· ···················· ····················
Y 5283 hCoV-19/Nigeria/BO-CV277/2020|EPI_ISL_729961|2020- ···················· ···················· ····················· ···················· ···················· ····················
Y 5284 hCoV-19/Nigeria/BO-CV278/2020|EPI_ISL_729962|2020- ···················· ···················· ····················· ···················· ···················· ····················
Y 5285 hCoV-19/Nigeria/BO-CV280/2020|EPI_ISL_729964|2020- ···················· ···················· ····················· ···················· ···················· ····················
Y 5286 hCoV-19/Nigeria/OS-CV285/2020|EPI_ISL_729966|2020- ···················· ···················· ····················· ···················· ···················· ····················
Y 5287 hCoV-19/Nigeria/OS-CV286/2020|EPI_ISL_729967|2020- ···················· ···················· ····················· ···················· ···················· ····················
Y 5288 hCoV-19/Nigeria/OS-CV287/2020|EPI_ISL_729968|2020- ···················· ···················· ····················· ···················· ···················· ····················
Y 5289 hCoV-19/Nigeria/OS-CV288/2020|EPI_ISL_729969|2020- ···················· ···················· ···········T········· ···················· ···················· ····················
Y 5290 hCoV-19/Nigeria/OS-CV290/2020|EPI_ISL_729970|2020- ···················· ···················· ····················· ···················· ···················· ····················
Y 5291 hCoV-19/Nigeria/OS-CV293/2020|EPI_ISL_729972|2020- ···················· ···················· ····················· ···················· ···················· ····················
Y 5292 hCoV-19/Nigeria/OS-CV294/2020|EPI_ISL_729973|2020- ···················· ···················· ····················· ···················· ···················· ····················
Y 5293 hCoV-19/Nigeria/OS-CV295/2020|EPI_ISL_729974|2020- ···················· ···················· ····················· ···················· ···················· ····················
Y 5294 hCoV-19/Nigeria/OS-CV296/2020|EPI_ISL_729975|2020- ···················· ···················· ····················· ···················· ···················· ····················
Y 5295 hCoV-19/Nigeria/OS-CV297/2020|EPI_ISL_729976|2020- ···················· ···················· ····················· ···················· ···················· ····················
Y 5296 hCoV-19/Nigeria/OS-CV299/2020|EPI_ISL_729977|2020- ···················· ···················· ····················· ···················· ···················· ····················
Y 5297 hCoV-19/Nigeria/OS-CV300/2020|EPI_ISL_729978|2020- ···················· ···················· ····················· ···················· ···················· ····················
Y 5298 hCoV-19/Nigeria/OS-CV301/2020|EPI_ISL_729979|2020- ···················· ···················· ····················· ···················· ···················· ····················
Y 5299 hCoV-19/Nigeria/OS-CV302/2020|EPI_ISL_729980|2020- ···················· ···················· ····················· ···················· ···················· ····················
Y 5300 hCoV-19/Nigeria/OS-CV304/2020|EPI_ISL_729982|2020- ···················· ···················· ····················· ···················· ···················· ····················
Y 5301 hCoV-19/Nigeria/OS-CV307/2020|EPI_ISL_729985|2020- ···················· ···················· ····················· ···················· ···················· ····················
Y 5302 hCoV-19/Nigeria/AK-CV308/2020|EPI_ISL_729986|2020- ···················· ···················· ····················· ···················· ···················· ····················
Y 5303 hCoV-19/Nigeria/AK-CV313/2020|EPI_ISL_729987|2020- ···················· ···················· ····················· ···················· ···················· ····················
Y 5304 hCoV-19/Nigeria/AK-CV314/2020|EPI_ISL_729988|2020- ···················· ···················· ····················· ···················· ···················· ····················
Y 5305 hCoV-19/Nigeria/AK-CV317/2020|EPI_ISL_729989|2020- ···················· ···················· ····················· ···················· ···················· ····················
Y 5306 hCoV-19/Nigeria/AK-CV324/2020|EPI_ISL_729994|2020- ···················· ···················· ····················· ···················· ···················· ····················
Y 5307 hCoV-19/Nigeria/BO-CV326/2020|EPI_ISL_729995|2020- ···················· ···················· ····················· ···················· ···················· ····················
Y 5308 hCoV-19/Nigeria/BO-CV330/2020|EPI_ISL_729999|2020- ···················· ···················· ····················· ···················· ···················· ····················
Y 5309 hCoV-19/England/ALDP-C3CF6C/2020|EPI_ISL_730941|20 ···················· ···················· ····················· ···················· ···················· ····················
Y 5310 hCoV-19/England/ALDP-C3D235/2020|EPI_ISL_730942|20 ···················· ···················· ····················· ···················· ···················· ····················
Y 5311 hCoV-19/England/ALDP-C3C8F8/2020|EPI_ISL_730943|20 ···················· ···················· ····················· ···················· ···················· ····················
Y 5312 hCoV-19/England/ALDP-C3C1A2/2020|EPI_ISL_730944|20 ···················· ···················· ····················· ···················· ···················· ····················
Y 5313 hCoV-19/England/ALDP-C3C746/2020|EPI_ISL_730945|20 ···················· ···················· ····················· ···················· ···················· ····················
Y 5314 hCoV-19/England/ALDP-C3C4E5/2020|EPI_ISL_730946|20 ···················· ···················· ····················· ···················· ···················· ····················
Y 5315 hCoV-19/England/ALDP-C3C45E/2020|EPI_ISL_730947|20 ···················· ···················· ····················· ···················· ···················· ····················
Y 5316 hCoV-19/England/ALDP-C3A274/2020|EPI_ISL_730948|20 ···················· ···················· ····················· ···················· ···················· ····················
Y 5317 hCoV-19/England/ALDP-C3DAD3/2020|EPI_ISL_730949|20 ···················· ···················· ····················· ···················· ···················· ····················
Y 5318 hCoV-19/England/ALDP-C3DAF1/2020|EPI_ISL_730950|20 ···················· ···················· ····················· ···················· ···················· ····················
Y 5319 hCoV-19/England/ALDP-C3C39D/2020|EPI_ISL_730951|20 ···················· ···················· ····················· ···················· ···················· ····················
Y 5320 hCoV-19/England/ALDP-C3C940/2020|EPI_ISL_730952|20 ···················· ···················· ····················· ···················· ···················· ····················
Y 5321 hCoV-19/England/ALDP-C3D94F/2020|EPI_ISL_730953|20 ···················· ···················· ····················· ···················· ···················· ····················
Y 5322 hCoV-19/England/ALDP-C3C421/2020|EPI_ISL_730954|20 ···················· ···················· ····················· ···················· ···················· ····················
Y 5323 hCoV-19/England/ALDP-C3DB49/2020|EPI_ISL_730956|20 ···················· ···················· ····················· ···················· ···················· ····················
Y 5324 hCoV-19/England/ALDP-C3C44F/2020|EPI_ISL_730957|20 ···················· ···················· ····················· ···················· ···················· ····················
Y 5325 hCoV-19/England/ALDP-C3D068/2020|EPI_ISL_730961|20 ···················· ···················· ····················· ···················· ···················· ····················
Y 5326 hCoV-19/England/ALDP-C3BDBE/2020|EPI_ISL_730963|20 ···················· ···················· ····················· ···················· ···················· ····················
Y 5327 hCoV-19/England/ALDP-C3B9E7/2020|EPI_ISL_730964|20 ···················· ···················· ····················· ···················· ···················· ····················
Y 5328 hCoV-19/England/ALDP-C3CD35/2020|EPI_ISL_730965|20 ···················· ···················· ····················· ···················· ···················· ····················
Y 5329 hCoV-19/England/ALDP-C39EDB/2020|EPI_ISL_730966|20 ···················· ···················· ····················· ···················· ···················· ····················
Y 5330 hCoV-19/England/ALDP-C3B6FF/2020|EPI_ISL_730968|20 ···················· ···················· ····················· ···················· ···················· ····················
Y 5331 hCoV-19/England/ALDP-C3A362/2020|EPI_ISL_730971|20 ···················· ···················· ····················· ···················· ···················· ····················
Y 5332 hCoV-19/England/ALDP-C3C667/2020|EPI_ISL_730972|20 ···················· ···················· ····················· ···················· ···················· ····················
Y 5333 hCoV-19/England/ALDP-C3D4F3/2020|EPI_ISL_730981|20 ···················· ···················· ····················· ···················· ···················· ····················
Y 5334 hCoV-19/England/ALDP-C3C8CB/2020|EPI_ISL_730982|20 ···················· ···················· ····················· ···················· ···················· ····················
Y 5335 hCoV-19/England/ALDP-C3C3E8/2020|EPI_ISL_730984|20 ···················· ···················· ····················· ···················· ···················· ····················
Y 5336 hCoV-19/England/ALDP-C3C834/2020|EPI_ISL_730985|20 ···················· ···················· ····················· ···················· ···················· ····················
Y 5337 hCoV-19/England/ALDP-C3A2DE/2020|EPI_ISL_730989|20 ···················· ···················· ····················· ···················· ···················· ····················
Y 5338 hCoV-19/England/ALDP-C3C0F0/2020|EPI_ISL_730990|20 ···················· ···················· ····················· ···················· ···················· ····················
Y 5339 hCoV-19/England/ALDP-C3C49A/2020|EPI_ISL_730991|20 ···················· ···················· ····················· ···················· ···················· ····················
Y 5340 hCoV-19/England/ALDP-C3BBE1/2020|EPI_ISL_730992|20 ···················· ···················· ····················· ···················· ···················· ····················
Y 5341 hCoV-19/England/ALDP-C3A2FC/2020|EPI_ISL_730994|20 ···················· ···················· ····················· ···················· ···················· ····················
Y 5342 hCoV-19/England/ALDP-C3DB0D/2020|EPI_ISL_730995|20 ···················· ···················· ····················· ···················· ···················· ····················
Y 5343 hCoV-19/England/ALDP-C3D4E4/2020|EPI_ISL_730996|20 ···················· ···················· ····················· ···················· ···················· ····················
Y 5344 hCoV-19/England/ALDP-C3CAE3/2020|EPI_ISL_730997|20 ···················· ···················· ····················· ···················· ···················· ····················
Y 5345 hCoV-19/England/ALDP-C3C333/2020|EPI_ISL_730998|20 ···················· ·A·················· ····················· ···················· ···················· ···A················
Y 5346 hCoV-19/England/ALDP-C3B738/2020|EPI_ISL_730999|20 ···················· ···················· ····················· ···················· ···················· ····················
Y 5347 hCoV-19/Uganda/UG089/2020|EPI_ISL_738000|2020-08-2 ···················· ···················· ····················· ···················· ···················· ····················
Y 5348 hCoV-19/Uganda/UG090/2020|EPI_ISL_738001|2020-08-1 ···················· ···················· ····················· ···················· ···················· ····················
Y 5349 hCoV-19/Uganda/UG094/2020|EPI_ISL_738002|2020-08-1 ···················· ···················· ····················· ···················· ···T················ ····················
Y 5350 hCoV-19/Uganda/UG095/2020|EPI_ISL_738003|2020-08-2 ···················· ···················· ····················· ···················· ···················· ····················
Y 5351 hCoV-19/Uganda/UG096/2020|EPI_ISL_738004|2020-08-2 ···················· ···················· ····················· ···················· ···················· ····················
Y 5352 hCoV-19/Uganda/UG097/2020|EPI_ISL_738005|2020-08-1 ···················· ···················· ····················· ···················· ···················· ····················
Y 5353 hCoV-19/Uganda/UG098/2020|EPI_ISL_738006|2020-08-2 ···················· ···················· ····················· ···················· ···················· ····················

| | | | | | | | |
|---|---|---|---|---|---|---|---|
| Y | 5354 hCoV-19/Uganda/UG099/2020\|EPI_ISL_738007\|2020-08-1 | .................... | .................... | .................... | .................... | .................... | .................... |
| Y | 5355 hCoV-19/Uganda/UG101/2020\|EPI_ISL_738008\|2020-08-1 | .................... | .................... | .................... | .................... | .................... | .................... |
| Y | 5356 hCoV-19/Uganda/UG102/2020\|EPI_ISL_738009\|2020-07-0 | .................... | .................... | .................... | .................... | .................... | .................... |
| Y | 5357 hCoV-19/Uganda/UG103/2020\|EPI_ISL_738010\|2020-07-0 | .................... | .................... | .................... | .................... | .................... | .................... |
| Y | 5358 hCoV-19/Uganda/UG104/2020\|EPI_ISL_738011\|2020-07-0 | .................... | .................... | .................... | .................... | .................... | .................... |
| Y | 5359 hCoV-19/Uganda/UG105/2020\|EPI_ISL_738012\|2020-07-0 | .................... | .................... | .................... | .................... | .................... | .................... |
| Y | 5360 hCoV-19/Uganda/UG106/2020\|EPI_ISL_738013\|2020-08-0 | .................... | .................... | .................... | .................... | .................... | .................... |
| Y | 5361 hCoV-19/Uganda/UG107/2020\|EPI_ISL_738014\|2020-08-0 | .................... | .................... | .................... | .................... | .................... | .................... |
| Y | 5362 hCoV-19/Uganda/UG108/2020\|EPI_ISL_738015\|2020-08-0 | .................... | .................... | .................... | .................... | .................... | .................... |
| Y | 5363 hCoV-19/Uganda/UG109/2020\|EPI_ISL_738016\|2020-08-1 | .................... | .................... | .................... | .................... | .................... | .................... |
| Y | 5364 hCoV-19/Uganda/UG110/2020\|EPI_ISL_738017\|2020-08-1 | .................... | .................... | .................... | .................... | .................... | .................... |
| Y | 5365 hCoV-19/Uganda/UG111/2020\|EPI_ISL_738018\|2020-08-1 | .................... | .................... | .................... | .................... | .................... | .................... |
| Y | 5366 hCoV-19/Uganda/UG112/2020\|EPI_ISL_738019\|2020-08-1 | .................... | .................... | .................... | .................... | .................... | .................... |
| Y | 5367 hCoV-19/Uganda/UG113/2020\|EPI_ISL_738020\|2020-08-1 | .................... | .................... | .................... | .................... | .................... | .................... |
| Y | 5368 hCoV-19/Uganda/UG114/2020\|EPI_ISL_738021\|2020-08-1 | .................... | .................... | .................... | .................... | .................... | .................... |
| Y | 5369 hCoV-19/Uganda/UG115/2020\|EPI_ISL_738022\|2020-08-2 | .................... | .................... | .................... | .................... | .................... | .................... |
| Y | 5370 hCoV-19/Uganda/UG116/2020\|EPI_ISL_738023\|2020-08-2 | .................... | .................... | .................... | .................... | .................... | .................... |
| Y | 5371 hCoV-19/Uganda/UG117/2020\|EPI_ISL_738024\|2020 | .................... | .................... | .................... | .................... | .................... | .................... |
| Y | 5372 hCoV-19/Uganda/UG118/2020\|EPI_ISL_738025\|2020-09-0 | .................... | .................... | .................... | .................... | .................... | .................... |
| Y | 5373 hCoV-19/Uganda/UG119/2020\|EPI_ISL_738026\|2020-09-0 | .................... | .................... | .................... | .................... | .................... | .................... |
| Y | 5374 hCoV-19/Uganda/UG120/2020\|EPI_ISL_738027\|2020-09-0 | .................... | .................... | .................... | .................... | .................... | .................... |
| Y | 5375 hCoV-19/Uganda/UG121/2020\|EPI_ISL_738028\|2020-09-0 | .................... | .................... | .................... | .................... | .................... | .................... |
| Y | 5376 hCoV-19/Uganda/UG122/2020\|EPI_ISL_738029\|2020-09-0 | .................... | .................... | .................... | .................... | .................... | .................... |
| Y | 5377 hCoV-19/Wales/PHWC-493FD4/2020\|EPI_ISL_726051\|2020 | .................... | .................... | .................... | .................... | .................... | .................... |
| Y | 5380 hCoV-19/Wales/PHWC-49400D/2020\|EPI_ISL_726054\|2020 | .................... | .................... | .................... | .................... | .................... | .................... |
| Y | 5381 hCoV-19/Wales/PHWC-49402B/2020\|EPI_ISL_726056\|2020 | .................... | .................... | .................... | .................... | .................... | .................... |
| Y | 5383 hCoV-19/Wales/PHWC-494049/2020\|EPI_ISL_726058\|2020 | .................... | .................... | .................... | .................... | .................... | .................... |
| Y | 5384 hCoV-19/Wales/PHWC-494058/2020\|EPI_ISL_726059\|2020 | .................... | .................... | .................... | .................... | .................... | .................... |
| Y | 5385 hCoV-19/Wales/PHWC-494076/2020\|EPI_ISL_726061\|2020 | .................... | .................... | .................... | .................... | .................... | .................... |
| Y | 5386 hCoV-19/Wales/PHWC-494085/2020\|EPI_ISL_726062\|2020 | .................... | .................... | .................... | .................... | .................... | .................... |
| Y | 5387 hCoV-19/Wales/PHWC-494094/2020\|EPI_ISL_726063\|2020 | .................... | .................... | .................... | .................... | .........NN......... | .................... |
| Y | 5388 hCoV-19/Wales/PHWC-4940D0/2020\|EPI_ISL_726066\|2020 | .................... | .................... | .................... | .................... | .................... | .................... |
| Y | 5389 hCoV-19/Wales/PHWC-4940EF/2020\|EPI_ISL_726067\|2020 | .................... | .................... | .................... | .................... | .................... | .................... |
| Y | 5390 hCoV-19/Wales/PHWC-4940FE/2020\|EPI_ISL_726068\|2020 | .................... | .................... | .................... | .................... | .................... | .................... |
| Y | 5391 hCoV-19/Wales/PHWC-49410A/2020\|EPI_ISL_726069\|2020 | .................... | .................... | .................... | .................... | .................... | .................... |
| Y | 5393 hCoV-19/Wales/PHWC-494128/2020\|EPI_ISL_726071\|2020 | .................... | .................... | .................... | .................... | .................... | .................... |
| Y | 5395 hCoV-19/Wales/PHWC-494146/2020\|EPI_ISL_726073\|2020 | .................... | .................... | .................... | .................... | .................... | .................... |
| Y | 5396 hCoV-19/Wales/PHWC-4941BF/2020\|EPI_ISL_726080\|2020 | .................... | .................... | .................... | .................... | .................... | .................... |
| Y | 5397 hCoV-19/Wales/PHWC-4941CE/2020\|EPI_ISL_726081\|2020 | .................... | .................... | .................... | .................... | .................... | .................... |
| Y | 5398 hCoV-19/Wales/PHWC-4941DD/2020\|EPI_ISL_726082\|2020 | .................... | .................... | .................... | .................... | .................... | .................... |
| Y | 5400 hCoV-19/Wales/PHWC-494216/2020\|EPI_ISL_726085\|2020 | .................... | .................... | .................... | .................... | .................... | .................... |
| Y | 5401 hCoV-19/Wales/PHWC-494225/2020\|EPI_ISL_726086\|2020 | .................... | .................... | .................... | .................... | .................... | .................... |
| Y | 5402 hCoV-19/Wales/PHWC-494234/2020\|EPI_ISL_726087\|2020 | .................... | .................... | .................... | .................... | .................... | .................... |
| Y | 5403 hCoV-19/Wales/PHWC-494243/2020\|EPI_ISL_726088\|2020 | .................... | .................... | .................... | .................... | .................... | .................... |
| Y | 5404 hCoV-19/Wales/PHWC-494252/2020\|EPI_ISL_726089\|2020 | .................... | .................... | .................... | .................... | .................... | .................... |
| Y | 5405 hCoV-19/Wales/PHWC-494261/2020\|EPI_ISL_726090\|2020 | .................... | .................... | .................... | .................... | .................... | .................... |
| Y | 5406 hCoV-19/Wales/PHWC-494270/2020\|EPI_ISL_726091\|2020 | .................... | .................... | .................... | .................... | .................... | .................... |
| Y | 5407 hCoV-19/Wales/PHWC-49428F/2020\|EPI_ISL_726092\|2020 | .................... | .................... | .................... | .................... | .................... | .................... |
| Y | 5409 hCoV-19/Wales/PHWC-4942BC/2020\|EPI_ISL_726095\|2020 | .................... | .................... | .................... | .................... | .................... | .................... |
| Y | 5410 hCoV-19/Wales/PHWC-4942CB/2020\|EPI_ISL_726096\|2020 | .................... | .................... | .................... | .................... | .................... | .................... |
| Y | 5411 hCoV-19/Wales/PHWC-4942DA/2020\|EPI_ISL_726097\|2020 | .................... | .................... | .................... | .................... | .................... | .................... |
| Y | 5412 hCoV-19/Wales/PHWC-4942F8/2020\|EPI_ISL_726098\|2020 | .................... | .................... | .................... | .................... | .................... | .................... |
| Y | 5414 hCoV-19/Wales/PHWC-499949/2020\|EPI_ISL_727380\|2020 | .................... | .................... | .................... | .................... | .................... | .................... |
| Y | 5417 hCoV-19/Wales/PHWC-4999A3/2020\|EPI_ISL_727383\|2020 | .................... | .................... | .................... | .................... | .................... | .................... |
| Y | 5418 hCoV-19/Wales/PHWC-4999B2/2020\|EPI_ISL_727384\|2020 | .................... | .................... | .................... | .................... | .................... | .................... |
| Y | 5420 hCoV-19/Wales/PHWC-4999EF/2020\|EPI_ISL_727387\|2020 | .................... | .................... | .................... | .................... | .................... | .................... |
| Y | 5421 hCoV-19/Wales/PHWC-4999FE/2020\|EPI_ISL_727388\|2020 | .................... | .................... | .................... | .................... | .................... | .................... |
| Y | 5423 hCoV-19/Wales/PHWC-499A28/2020\|EPI_ISL_727390\|2020 | .................... | .................... | .................... | .................... | .................... | .................... |
| Y | 5426 hCoV-19/Wales/PHWC-499A55/2020\|EPI_ISL_727393\|2020 | .................... | .................... | .................... | .................... | .................... | .................... |
| Y | 5427 hCoV-19/Wales/PHWC-499A73/2020\|EPI_ISL_727395\|2020 | .................... | .................... | .................... | .................... | .................... | .................... |
| Y | 5430 hCoV-19/Wales/PHWC-499ABF/2020\|EPI_ISL_727399\|2020 | .................... | .................... | .................... | .................... | .................... | .................... |
| Y | 5431 hCoV-19/Netherlands/NH-RIVM-20485/2020\|EPI_ISL_728 | .................... | .................... | .................... | .................... | .................... | .................... |
| Y | 5432 hCoV-19/Netherlands/NH-RIVM-20486/2020\|EPI_ISL_728 | .................... | .................... | .................... | .................... | .....T.............. | .................... |
| Y | 5433 hCoV-19/Netherlands/NH-RIVM-20487/2020\|EPI_ISL_728 | .................... | .................... | .................... | .................... | .................... | .................... |
| Y | 5434 hCoV-19/Netherlands/LI-RIVM-20488/2020\|EPI_ISL_728 | .................... | .................... | .................... | .................... | .................... | .................... |
| Y | 5435 hCoV-19/Netherlands/ZH-RIVM-20489/2020\|EPI_ISL_728 | .................... | .................... | .................... | .................... | .................... | .................... |
| Y | 5436 hCoV-19/Netherlands/ZH-RIVM-20490/2020\|EPI_ISL_728 | .................... | .................... | .................... | .................... | .................... | .................... |
| Y | 5437 hCoV-19/Netherlands/ZH-RIVM-20491/2020\|EPI_ISL_728 | .................... | .................... | .................... | .................... | .................... | .................... |
| Y | 5438 hCoV-19/Netherlands/ZH-RIVM-20492/2020\|EPI_ISL_728 | .................... | .................... | .................... | .................... | .................... | .................... |
| Y | 5439 hCoV-19/Russia/AST-RII-MH4459S/2020\|EPI_ISL_733200 | .................... | .................... | .................... | .................... | .................... | .................... |
| Y | 5440 hCoV-19/Russia/AST-RII-MH4465S/2020\|EPI_ISL_733203 | .................... | .................... | .................... | .................... | .................... | .................... |
| Y | 5441 hCoV-19/Russia/AST-RII-MH4466S/2020\|EPI_ISL_733204 | .................... | .................... | .................... | .................... | .................... | .................... |
| Y | 5442 hCoV-19/Russia/AST-RII-MH4468S/2020\|EPI_ISL_733205 | .................... | .................... | .................... | .................... | .................... | .................... |
| Y | 5443 hCoV-19/Russia/AST-RII-MH4470S/2020\|EPI_ISL_733206 | .................... | .................... | .................... | .................... | .................... | .................... |
| Y | 5445 hCoV-19/Russia/AST-RII-MH4476S/2020\|EPI_ISL_733208 | .................... | .................... | .................... | .................... | .................... | .................... |
| Y | 5446 hCoV-19/Russia/AST-RII-MH4490S/2020\|EPI_ISL_733210 | .................... | .................... | .................... | .................... | .................... | .................... |
| Y | 5447 hCoV-19/Russia/AST-RII-MH4494S/2020\|EPI_ISL_733211 | .................... | .................... | .................... | .................... | .................... | .................... |
| Y | 5448 hCoV-19/Russia/AST-RII-MH4571S/2020\|EPI_ISL_733215 | .................... | .................... | .................... | .................... | .................... | .................... |
| Y | 5449 hCoV-19/Russia/AST-RII-MH4597S/2020\|EPI_ISL_733220 | .................... | .................... | .................... | .................... | .................... | .................... |
| Y | 5450 hCoV-19/Russia/AST-RII-MH4609S/2020\|EPI_ISL_733222 | .................... | .................... | .................... | .................... | .................... | .................... |

| | | | | | | | | |
|---|---|---|---|---|---|---|---|---|
| Y | 5451 | hCoV-19/Russia/SVE-RII-MH4633S/2020\|EPI_ISL_733223 | ···················· | ···················· | ····················· | ···················· | ···················· | ···················· |
| Y | 5452 | hCoV-19/Russia/SVE-RII-MH4660S/2020\|EPI_ISL_733224 | ···················· | ···················· | ····················· | ···················· | ···················· | ···················· |
| Y | 5453 | hCoV-19/Russia/SVE-RII-MH4662S/2020\|EPI_ISL_733225 | ···················· | ···················· | ····················· | ···················· | ···················· | ···················· |
| Y | 5454 | hCoV-19/Russia/SVE-RII-MH4663S/2020\|EPI_ISL_733226 | ···················· | ···················· | ····················· | ···················· | ···················· | ···················· |
| Y | 5455 | hCoV-19/Russia/SVE-RII-MH4667S/2020\|EPI_ISL_733228 | ···················· | ···················· | ····················· | ···················· | ···················· | ···················· |
| Y | 5456 | hCoV-19/Russia/SVE-RII-MH4673S/2020\|EPI_ISL_733231 | ···················· | ···················· | ····················· | ···················· | ···················· | ···················· |
| Y | 5457 | hCoV-19/Russia/SVE-RII-MH4675S/2020\|EPI_ISL_733232 | ···················· | ···················· | ····················· | ···················· | ···················· | ···················· |
| Y | 5458 | hCoV-19/Russia/SVE-RII-MH4681S/2020\|EPI_ISL_733233 | ···················· | ···················· | ····················· | ···················· | ···················· | ···················· |
| Y | 5460 | hCoV-19/Russia/SVE-RII-MH4700S/2020\|EPI_ISL_733240 | ···················· | ···················· | ····················· | ···················· | ···················· | ···················· |
| Y | 5461 | hCoV-19/Russia/SAR-RII-MH4702S/2020\|EPI_ISL_733241 | ···················· | ···················· | ····················· | ···················· | ···················· | ···················· |
| Y | 5462 | hCoV-19/Russia/KDA-RII-MH4710S/2020\|EPI_ISL_733243 | ···················· | ···················· | ····················· | ···················· | ···················· | ···················· |
| Y | 5463 | hCoV-19/Russia/KDA-RII-MH4721S/2020\|EPI_ISL_733246 | ···················· | ···················· | ····················· | ···················· | ···················· | ···················· |
| Y | 5464 | hCoV-19/Russia/KDA-RII-MH4727S/2020\|EPI_ISL_733247 | ···················· | ···················· | ····················· | ···················· | ···················· | ···················· |
| Y | 5465 | hCoV-19/Russia/KDA-RII-MH4730S/2020\|EPI_ISL_733248 | ···················· | ···················· | ····················· | ···················· | ···················· | ···················· |
| Y | 5466 | hCoV-19/Russia/KDA-RII-MH4731S/2020\|EPI_ISL_733249 | ···················· | ···················· | ····················· | ···················· | ···················· | ···················· |
| Y | 5467 | hCoV-19/Russia/KDA-RII-MH4740S/2020\|EPI_ISL_733252 | ···················· | ···················· | ····················· | ···················· | ···················· | ···················· |
| Y | 5468 | hCoV-19/Russia/KDA-RII-MH4744S/2020\|EPI_ISL_733254 | ···················· | ···················· | ····················· | ···················· | ···················· | ···················· |
| Y | 5469 | hCoV-19/Russia/KDA-RII-MH4745S/2020\|EPI_ISL_733255 | ···················· | ···················· | ····················· | ···················· | ···················· | ···················· |
| Y | 5470 | hCoV-19/Russia/KDA-RII-MH4747S/2020\|EPI_ISL_733256 | ···················· | ···················· | ·········T··········· | ···················· | ···················· | ···················· |
| Y | 5471 | hCoV-19/Russia/KDA-RII-MH4836S/2020\|EPI_ISL_733269 | ···················· | ···················· | ····················· | ···················· | ···················· | ···················· |
| Y | 5472 | hCoV-19/Russia/KDA-RII-MH4846S/2020\|EPI_ISL_733272 | ···················· | ···················· | ····················· | ···················· | ···················· | ···················· |
| Y | 5473 | hCoV-19/Russia/KDA-RII-MH4855S/2020\|EPI_ISL_733275 | ···················· | ···················· | ····················· | ···················· | ···················· | ···················· |
| Y | 5474 | hCoV-19/Russia/KDA-RII-MH4868S/2020\|EPI_ISL_733277 | ···················· | ···················· | ····················· | ···················· | ···················· | ···················· |
| Y | 5475 | hCoV-19/Russia/KDA-RII-MH4871S/2020\|EPI_ISL_733278 | ···················· | ···················· | ····················· | ···················· | ···················· | ···················· |
| Y | 5476 | hCoV-19/Russia/KDA-RII-MH4875S/2020\|EPI_ISL_733279 | ···················· | ···················· | ····················· | ···················· | ···················· | ···················· |
| Y | 5477 | hCoV-19/Russia/KDA-RII-MH4887S/2020\|EPI_ISL_733280 | ···················· | ···················· | ····················· | ···················· | ···················· | ···················· |
| Y | 5480 | hCoV-19/Russia/KDA-RII-MH4928S/2020\|EPI_ISL_733287 | ···················· | ···A················ | ····················· | ···················· | ···················· | ···················· |
| Y | 5481 | hCoV-19/Russia/KDA-RII-MH4936S/2020\|EPI_ISL_733289 | ···················· | ···················· | ····················· | ···················· | ···················· | ···················· |
| Y | 5483 | hCoV-19/Belgium/rega-03031132/2020\|EPI_ISL_734503\| | ···················· | ···················· | ····················· | ···················· | ···················· | ···················· |
| Y | 5484 | hCoV-19/Belgium/rega-03121133/2020\|EPI_ISL_734504\| | ···················· | ···················· | ····················· | ···················· | ···················· | ···················· |
| Y | 5485 | hCoV-19/Belgium/rega-0315855/2020\|EPI_ISL_734505\|2 | ···················· | ·A·················· | ····················· | ···················· | ···················· | ···················· |
| Y | 5486 | hCoV-19/Belgium/rega-0317857/2020\|EPI_ISL_734506\|2 | ···················· | ···················· | ····················· | ···················· | ···················· | ···················· |
| Y | 5487 | hCoV-19/Belgium/rega-0319858/2020\|EPI_ISL_734507\|2 | ···················· | ···················· | ····················· | ···················· | ···················· | ···················· |
| Y | 5488 | hCoV-19/Belgium/rega-0319859/2020\|EPI_ISL_734508\|2 | ···················· | ···················· | ····················· | ···················· | ···················· | ···················· |
| Y | 5489 | hCoV-19/Belgium/rega-0319860/2020\|EPI_ISL_734509\|2 | ···················· | ···················· | ····················· | ···················· | ···················· | ···················· |
| Y | 5490 | hCoV-19/Belgium/rega-0320854/2020\|EPI_ISL_734510\|2 | ···················· | ···················· | ····················· | ···················· | ···················· | ···················· |
| Y | 5491 | hCoV-19/Belgium/rega-0320861/2020\|EPI_ISL_734511\|2 | ···················· | ···················· | ····················· | ···················· | ···················· | ···················· |
| Y | 5492 | hCoV-19/Belgium/rega-0320862/2020\|EPI_ISL_734512\|2 | ···················· | ···················· | ····················· | ···················· | ···················· | ···················· |
| Y | 5493 | hCoV-19/Belgium/rega-0320863/2020\|EPI_ISL_734513\|2 | ···················· | ···················· | ····················· | ···················· | ···················· | ···················· |
| Y | 5494 | hCoV-19/Belgium/rega-0322864/2020\|EPI_ISL_734514\|2 | ···················· | ···················· | ····················· | ···················· | ···················· | ···················· |
| Y | 5495 | hCoV-19/Belgium/rega-0322865/2020\|EPI_ISL_734515\|2 | ···················· | ···················· | ····················· | ···················· | ···················· | ···················· |
| Y | 5496 | hCoV-19/Belgium/rega-0322876/2020\|EPI_ISL_734516\|2 | ···················· | ···················· | ····················· | ···················· | ···················· | ···················· |
| Y | 5497 | hCoV-19/Belgium/rega-0323867/2020\|EPI_ISL_734517\|2 | ···················· | ···················· | ····················· | ···················· | ···················· | ···················· |
| Y | 5498 | hCoV-19/Belgium/rega-0323868/2020\|EPI_ISL_734518\|2 | ···················· | ···················· | ····················· | ···················· | ···················· | ···················· |
| Y | 5499 | hCoV-19/Belgium/rega-0323869/2020\|EPI_ISL_734519\|2 | ···················· | ···················· | ····················· | ···················· | ···················· | ···················· |
| Y | 5500 | hCoV-19/Belgium/rega-0324870/2020\|EPI_ISL_734520\|2 | ···················· | ···················· | ····················· | ···················· | ···················· | ···················· |
| Y | 5501 | hCoV-19/Belgium/rega-0325871/2020\|EPI_ISL_734521\|2 | ···················· | ···················· | ····················· | ···················· | ···················· | ···················· |
| Y | 5502 | hCoV-19/Belgium/rega-0325872/2020\|EPI_ISL_734522\|2 | ···················· | ···················· | ····················· | ···················· | ···················· | ···················· |
| Y | 5503 | hCoV-19/Belgium/rega-0326873/2020\|EPI_ISL_734523\|2 | ···················· | ···················· | ····················· | ···················· | ···················· | ···················· |
| Y | 5504 | hCoV-19/Belgium/rega-0328874/2020\|EPI_ISL_734524\|2 | ···················· | ···················· | ····················· | ···················· | ···················· | ···················· |
| Y | 5505 | hCoV-19/Belgium/rega-0330875/2020\|EPI_ISL_734525\|2 | ···················· | ···················· | ····················· | ···················· | ···················· | ···················· |
| Y | 5506 | hCoV-19/Belgium/rega-0331866/2020\|EPI_ISL_734526\|2 | ···················· | ···················· | ····················· | ···················· | ···················· | ···················· |
| Y | 5507 | hCoV-19/Belgium/rega-0331877/2020\|EPI_ISL_734527\|2 | ···················· | ···················· | ····················· | ···················· | ···················· | ···················· |
| Y | 5508 | hCoV-19/Belgium/rega-0401878/2020\|EPI_ISL_734528\|2 | ···················· | ···················· | ····················· | ···················· | ···················· | ···················· |
| Y | 5509 | hCoV-19/Belgium/rega-0401879/2020\|EPI_ISL_734529\|2 | ···················· | ·A·················· | ····················· | ···················· | ···················· | ···················· |
| Y | 5510 | hCoV-19/Belgium/rega-0401880/2020\|EPI_ISL_734530\|2 | ···················· | ···················· | ····················· | ···················· | ···················· | ···················· |
| Y | 5511 | hCoV-19/Belgium/rega-0402881/2020\|EPI_ISL_734531\|2 | ···················· | ···················· | ····················· | ···················· | ···················· | ···················· |
| Y | 5512 | hCoV-19/Belgium/rega-0403882/2020\|EPI_ISL_734532\|2 | ···················· | ···················· | ····················· | ···················· | ···················· | ···················· |
| Y | 5513 | hCoV-19/Belgium/rega-0403883/2020\|EPI_ISL_734533\|2 | ···················· | ···················· | ····················· | ···················· | ···················· | ···················· |
| Y | 5514 | hCoV-19/Belgium/rega-0403884/2020\|EPI_ISL_734534\|2 | ···················· | ···················· | ····················· | ···················· | ···················· | ···················· |
| Y | 5515 | hCoV-19/Belgium/rega-0404885/2020\|EPI_ISL_734535\|2 | ···················· | ···················· | ····················· | ···················· | ···················· | ···················· |
| Y | 5516 | hCoV-19/Belgium/rega-0406856/2020\|EPI_ISL_734536\|2 | ···················· | ···················· | ····················· | ···················· | ···················· | ···················· |
| Y | 5517 | hCoV-19/Belgium/rega-0406886/2020\|EPI_ISL_734537\|2 | ···················· | ···················· | ····················· | ···················· | ···················· | ···················· |
| Y | 5518 | hCoV-19/Belgium/rega-0406887/2020\|EPI_ISL_734538\|2 | ···················· | ···················· | ····················· | ···················· | ···················· | ···················· |
| Y | 5519 | hCoV-19/Belgium/rega-0406888/2020\|EPI_ISL_734539\|2 | ···················· | ···················· | ····················· | ···················· | ···················· | ···················· |
| Y | 5520 | hCoV-19/Belgium/rega-0407889/2020\|EPI_ISL_734540\|2 | ···················· | ···················· | ····················· | ···················· | ···················· | ···················· |
| Y | 5521 | hCoV-19/Belgium/rega-0407890/2020\|EPI_ISL_734541\|2 | ···················· | ···················· | ····················· | ···················· | ···················· | ···················· |
| Y | 5522 | hCoV-19/Belgium/rega-0407891/2020\|EPI_ISL_734542\|2 | ···················· | ···················· | ····················· | ···················· | ···················· | ···················· |
| Y | 5523 | hCoV-19/Belgium/rega-0407892/2020\|EPI_ISL_734543\|2 | ···················· | ···················· | ····················· | ···················· | ···················· | ···················· |
| Y | 5524 | hCoV-19/Belgium/rega-0408689/2020\|EPI_ISL_734544\|2 | ···················· | ···················· | ····················· | ···················· | ···················· | ···················· |
| Y | 5525 | hCoV-19/Belgium/rega-0408893/2020\|EPI_ISL_734545\|2 | ···················· | ···················· | ····················· | ···················· | ···················· | ···················· |
| Y | 5526 | hCoV-19/Belgium/rega-0408894/2020\|EPI_ISL_734546\|2 | ···················· | ···················· | ····················· | ···················· | ···················· | ···················· |
| Y | 5527 | hCoV-19/Belgium/rega-0408895/2020\|EPI_ISL_734547\|2 | ···················· | ···················· | ····················· | ···················· | ···················· | ···················· |
| Y | 5528 | hCoV-19/Belgium/rega-0408896/2020\|EPI_ISL_734548\|2 | ···················· | ···················· | ····················· | ···················· | ···················· | ···················· |
| Y | 5529 | hCoV-19/Belgium/rega-0408897/2020\|EPI_ISL_734549\|2 | ···················· | ···················· | ····················· | ···················· | ···················· | ···················· |
| Y | 5530 | hCoV-19/Belgium/rega-0409560/2020\|EPI_ISL_734550\|2 | ···················· | ···················· | ····················· | ···················· | ···················· | ···················· |
| Y | 5531 | hCoV-19/Belgium/rega-0409561/2020\|EPI_ISL_734551\|2 | ···················· | ···················· | ····················· | ···················· | ···················· | ···················· |
| Y | 5532 | hCoV-19/Belgium/rega-0409562/2020\|EPI_ISL_734552\|2 | ···················· | ···················· | ····················· | ···················· | ···················· | ···················· |
| Y | 5533 | hCoV-19/Belgium/rega-0409563/2020\|EPI_ISL_734553\|2 | ···················· | ···················· | ····················· | ···················· | ···················· | ···················· |
| Y | 5534 | hCoV-19/Belgium/rega-0409564/2020\|EPI_ISL_734554\|2 | ···················· | ···················· | ····················· | ···················· | ···················· | ···················· |

| | | | | | | | |
|---|---|---|---|---|---|---|---|
| Y | 5535 hCoV-19/Belgium/rega-0409565/2020\|EPI_ISL_734555\|2 | ···················· | ···················· | ····················· | ···················· | ···················· | ···················· |
| Y | 5536 hCoV-19/Belgium/rega-0409566/2020\|EPI_ISL_734556\|2 | ···················· | ···················· | ····················· | ···················· | ···················· | ···················· |
| Y | 5537 hCoV-19/Belgium/rega-0409567/2020\|EPI_ISL_734557\|2 | ···················· | ···················· | ····················· | ···················· | ···················· | ···················· |
| Y | 5538 hCoV-19/Belgium/rega-0409568/2020\|EPI_ISL_734558\|2 | ···················· | ···················· | ····················· | ···················· | ···················· | ···················· |
| Y | 5539 hCoV-19/Belgium/rega-0409569/2020\|EPI_ISL_734559\|2 | ···················· | ···················· | ····················· | ···················· | ···················· | ···················· |
| Y | 5540 hCoV-19/Belgium/rega-0410570/2020\|EPI_ISL_734560\|2 | ···················· | ···················· | ····················· | ···················· | ···················· | ···················· |
| Y | 5541 hCoV-19/Belgium/rega-0410571/2020\|EPI_ISL_734561\|2 | ···················· | ···················· | ····················· | ···················· | ···················· | ···················· |
| Y | 5542 hCoV-19/Belgium/rega-0410572/2020\|EPI_ISL_734562\|2 | ···················· | ···················· | ····················· | ···················· | ···················· | ···················· |
| Y | 5543 hCoV-19/Belgium/rega-0410573/2020\|EPI_ISL_734563\|2 | ···················· | ···················· | ····················· | ···················· | ···················· | ···················· |
| Y | 5544 hCoV-19/Belgium/rega-0410574/2020\|EPI_ISL_734564\|2 | ···················· | ···················· | ····················· | ···················· | ···················· | ···················· |
| Y | 5545 hCoV-19/Belgium/rega-0411898/2020\|EPI_ISL_734565\|2 | ···················· | ···················· | ····················· | ···················· | ···················· | ···················· |
| Y | 5546 hCoV-19/Belgium/rega-0411899/2020\|EPI_ISL_734566\|2 | ···················· | ···················· | ····················· | ···················· | ···················· | ···················· |
| Y | 5547 hCoV-19/Belgium/rega-0412901/2020\|EPI_ISL_734567\|2 | ···················· | ···················· | ····················· | ···················· | ···················· | ···················· |
| Y | 5548 hCoV-19/Belgium/rega-0413576/2020\|EPI_ISL_734568\|2 | ···················· | ···················· | ····················· | ···················· | ···················· | ···················· |
| Y | 5549 hCoV-19/Belgium/rega-0413577/2020\|EPI_ISL_734569\|2 | ···················· | ···················· | ····················· | ···················· | ···················· | ···················· |
| Y | 5550 hCoV-19/Belgium/rega-0413578/2020\|EPI_ISL_734570\|2 | ···················· | ···················· | ····················· | ···················· | ···················· | ···················· |
| Y | 5551 hCoV-19/Belgium/rega-0413579/2020\|EPI_ISL_734571\|2 | ···················· | ···················· | ····················· | ···················· | ···················· | ···················· |
| Y | 5552 hCoV-19/Belgium/rega-0414489/2020\|EPI_ISL_734572\|2 | ···················· | ···················· | ····················· | ···················· | ···················· | ···················· |
| Y | 5553 hCoV-19/Belgium/rega-0414490/2020\|EPI_ISL_734573\|2 | ···················· | ···················· | ····················· | ···················· | ···················· | ···················· |
| Y | 5554 hCoV-19/Belgium/rega-0414491/2020\|EPI_ISL_734574\|2 | ···················· | ···················· | ····················· | ···················· | ···················· | ···················· |
| Y | 5555 hCoV-19/Belgium/rega-0414492/2020\|EPI_ISL_734575\|2 | ···················· | ···················· | ····················· | ···················· | ···················· | ···················· |
| Y | 5556 hCoV-19/Belgium/rega-0414493/2020\|EPI_ISL_734576\|2 | ···················· | ···················· | ····················· | ···················· | ···················· | ···················· |
| Y | 5557 hCoV-19/Belgium/rega-0414494/2020\|EPI_ISL_734577\|2 | ···················· | ···················· | ····················· | ···················· | ···················· | ···················· |
| Y | 5558 hCoV-19/Belgium/rega-0414495/2020\|EPI_ISL_734578\|2 | ···················· | ···················· | ····················· | ···················· | ···················· | ···················· |
| Y | 5559 hCoV-19/Belgium/rega-0414580/2020\|EPI_ISL_734579\|2 | ···················· | ···················· | ····················· | ···················· | ···················· | ···················· |
| Y | 5560 hCoV-19/Belgium/rega-0414582/2020\|EPI_ISL_734580\|2 | ···················· | ···················· | ····················· | ···················· | ···················· | ···················· |
| Y | 5561 hCoV-19/Belgium/rega-0414583/2020\|EPI_ISL_734581\|2 | ···················· | ···················· | ····················· | ···················· | ···················· | ···················· |
| Y | 5562 hCoV-19/Belgium/rega-0414584/2020\|EPI_ISL_734582\|2 | ···················· | ···················· | ····················· | ···················· | ···················· | ···················· |
| Y | 5563 hCoV-19/Belgium/rega-0414902/2020\|EPI_ISL_734583\|2 | ···················· | ···················· | ····················· | ···················· | ···················· | ···················· |
| Y | 5564 hCoV-19/Belgium/rega-0414903/2020\|EPI_ISL_734584\|2 | ···················· | ···················· | ····················· | ···················· | ···················· | ···················· |
| Y | 5565 hCoV-19/Belgium/rega-0414904/2020\|EPI_ISL_734585\|2 | ···················· | ···················· | ····················· | ···················· | ···················· | ···················· |
| Y | 5566 hCoV-19/Belgium/rega-0415905/2020\|EPI_ISL_734586\|2 | ···················· | ···················· | ····················· | ···················· | ···················· | ···················· |
| Y | 5567 hCoV-19/Belgium/rega-0415906/2020\|EPI_ISL_734587\|2 | ···················· | ···················· | ····················· | ···················· | ···················· | ···················· |
| Y | 5568 hCoV-19/Belgium/rega-0416496/2020\|EPI_ISL_734588\|2 | ···················· | ···················· | ····················· | ···················· | ···················· | ···················· |
| Y | 5569 hCoV-19/Belgium/rega-0416497/2020\|EPI_ISL_734589\|2 | ···················· | ···················· | ····················· | ···················· | ···················· | ···················· |
| Y | 5570 hCoV-19/Belgium/rega-0416500/2020\|EPI_ISL_734592\|2 | ···················· | ···················· | ····················· | ···················· | ···················· | ···················· |
| Y | 5571 hCoV-19/Belgium/rega-0416501/2020\|EPI_ISL_734593\|2 | ···················· | ···················· | ····················· | ···················· | ···················· | ···················· |
| Y | 5572 hCoV-19/Belgium/rega-0416502/2020\|EPI_ISL_734594\|2 | ···················· | ···················· | ····················· | ···················· | ···················· | ···················· |
| Y | 5573 hCoV-19/Belgium/rega-0416907/2020\|EPI_ISL_734595\|2 | ···················· | ···················· | ····················· | ···················· | ···················· | ···················· |
| Y | 5574 hCoV-19/Belgium/rega-0417503/2020\|EPI_ISL_734596\|2 | ···················· | ···················· | ····················· | ···················· | ···················· | ···················· |
| Y | 5575 hCoV-19/Belgium/rega-0417504/2020\|EPI_ISL_734597\|2 | ···················· | ···················· | ····················· | ···················· | ···················· | ···················· |
| Y | 5576 hCoV-19/Belgium/rega-0417505/2020\|EPI_ISL_734598\|2 | ···················· | ···················· | ····················· | ···················· | ···················· | ···················· |
| Y | 5577 hCoV-19/Belgium/rega-0417507/2020\|EPI_ISL_734599\|2 | ···················· | ···················· | ····················· | ···················· | ···················· | ···················· |
| Y | 5578 hCoV-19/England/MILK-C6397D/2020\|EPI_ISL_735802\|20 | ···················· | ···················· | ····················· | ···················· | ···················· | ···················· |
| Y | 5579 hCoV-19/England/MILK-C62264/2020\|EPI_ISL_735804\|20 | ···················· | ···················· | ····················· | ···················· | ···················· | ···················· |
| Y | 5580 hCoV-19/England/MILK-C61317/2020\|EPI_ISL_735806\|20 | ···················· | ···················· | ····················· | ···················· | ···················· | ···················· |
| Y | 5581 hCoV-19/England/MILK-C61F04/2020\|EPI_ISL_735807\|20 | ···················· | ···················· | ····················· | ···················· | ···················· | ···················· |
| Y | 5582 hCoV-19/England/MILK-C6112C/2020\|EPI_ISL_735812\|20 | ···················· | ···················· | ····················· | ···················· | ···················· | ···················· |
| Y | 5583 hCoV-19/England/MILK-C63B0E/2020\|EPI_ISL_735814\|20 | ···················· | ···················· | ····················· | ···················· | ···················· | ···················· |
| Y | 5584 hCoV-19/England/MILK-C6388F/2020\|EPI_ISL_735817\|20 | ···················· | ···················· | ····················· | ···················· | ···················· | ···················· |
| Y | 5585 hCoV-19/England/MILK-C61EBC/2020\|EPI_ISL_735833\|20 | ···················· | ···················· | ····················· | ···················· | ···················· | ···················· |
| Y | 5586 hCoV-19/England/MILK-C62088/2020\|EPI_ISL_735836\|20 | ···················· | ···················· | ····················· | ···················· | ···················· | ···················· |
| Y | 5587 hCoV-19/England/MILK-C61BD3/2020\|EPI_ISL_735837\|20 | ···················· | ·A·················· | ····················· | ···················· | ···················· | ···················· |
| Y | 5588 hCoV-19/England/MILK-C6204C/2020\|EPI_ISL_735838\|20 | ···················· | ···················· | ····················· | ···················· | ···················· | ···················· |
| Y | 5589 hCoV-19/England/MILK-C615C6/2020\|EPI_ISL_735844\|20 | ···················· | ···················· | ····················· | ···················· | ···················· | ···················· |
| Y | 5590 hCoV-19/England/MILK-C6172A/2020\|EPI_ISL_735846\|20 | ···················· | ···················· | ····················· | ···················· | ···················· | ···················· |
| Y | 5591 hCoV-19/England/MILK-C61872/2020\|EPI_ISL_735847\|20 | ···················· | ···················· | ····················· | ···················· | ······T············· | ···················· |
| Y | 5592 hCoV-19/England/MILK-C62167/2020\|EPI_ISL_735855\|20 | ···················· | ···················· | ····················· | ···················· | ···················· | ···················· |
| Y | 5593 hCoV-19/England/MILK-C61854/2020\|EPI_ISL_735858\|20 | ···················· | ···················· | ····················· | ···················· | ···················· | ···················· |
| Y | 5594 hCoV-19/England/QEUH-C5D554/2020\|EPI_ISL_735861\|20 | ···················· | ·A·················· | ····················· | ···················· | ···················· | ···················· |
| Y | 5595 hCoV-19/England/ALDP-C59190/2020\|EPI_ISL_735862\|20 | ···················· | ···················· | ····················· | ···················· | ···················· | ···················· |
| Y | 5596 hCoV-19/England/ALDP-C58146/2020\|EPI_ISL_735863\|20 | ···················· | ···················· | ····················· | ···················· | ···················· | ···················· |
| Y | 5597 hCoV-19/England/ALDP-C58C90/2020\|EPI_ISL_735868\|20 | ···················· | ···················· | ····················· | ···················· | ···················· | ···················· |
| Y | 5598 hCoV-19/England/ALDP-C59FE1/2020\|EPI_ISL_735871\|20 | ···················· | ···················· | ····················· | ···················· | ···················· | ···················· |
| Y | 5599 hCoV-19/England/ALDP-C58656/2020\|EPI_ISL_735872\|20 | ···················· | ···················· | ····················· | ···················· | ···················· | ···················· |
| Y | 5600 hCoV-19/England/ALDP-C59707/2020\|EPI_ISL_735875\|20 | ···················· | ···················· | ····················· | ···················· | ···················· | ···················· |
| Y | 5601 hCoV-19/Wales/ALDP-C586B0/2020\|EPI_ISL_735876\|2020 | ···················· | ···················· | ····················· | ···················· | ···················· | ···················· |
| Y | 5602 hCoV-19/England/ALDP-C57C82/2020\|EPI_ISL_735877\|20 | ···················· | ···················· | ····················· | ···················· | ···················· | ···················· |
| Y | 5603 hCoV-19/England/ALDP-C584F2/2020\|EPI_ISL_735881\|20 | ···················· | ···················· | ····················· | ···················· | ···················· | ···················· |
| Y | 5604 hCoV-19/England/ALDP-C58EF4/2020\|EPI_ISL_735882\|20 | ···················· | ···················· | ····················· | ···················· | ···················· | ···················· |
| Y | 5605 hCoV-19/England/ALDP-C59734/2020\|EPI_ISL_735883\|20 | ···················· | ···················· | ····················· | ···················· | ······T············· | ···················· |
| Y | 5606 hCoV-19/England/ALDP-C5886F/2020\|EPI_ISL_735887\|20 | ·····G·············· | ···················· | ····················· | ···················· | ···················· | ···················· |
| Y | 5607 hCoV-19/England/ALDP-C585B3/2020\|EPI_ISL_735888\|20 | ···················· | ···················· | ····················· | ···················· | ···················· | ···················· |
| Y | 5608 hCoV-19/England/ALDP-C57FE3/2020\|EPI_ISL_735889\|20 | ···················· | ···················· | ····················· | ···················· | ···················· | ···················· |
| Y | 5609 hCoV-19/England/ALDP-C580D0/2020\|EPI_ISL_735890\|20 | ···················· | ···················· | ····················· | ···················· | ···················· | ···················· |
| Y | 5610 hCoV-19/England/ALDP-C57D70/2020\|EPI_ISL_735891\|20 | ···················· | ···················· | ····················· | ···················· | ···················· | ···················· |
| Y | 5611 hCoV-19/England/QEUH-C5BCBB/2020\|EPI_ISL_735894\|20 | ···················· | ···················· | ····················· | ···················· | ···················· | ···················· |
| Y | 5612 hCoV-19/England/ALDP-C5853B/2020\|EPI_ISL_735897\|20 | ···················· | ···················· | ····················· | ···················· | ···················· | ···················· |
| Y | 5613 hCoV-19/England/ALDP-C58BFD/2020\|EPI_ISL_735899\|20 | ···················· | ···················· | ····················· | ···················· | ···················· | ···················· |
| Y | 5614 hCoV-19/Uganda/UG123/2020\|EPI_ISL_738030\|2020-08-1 | ···················· | ···················· | ····················· | ···················· | ···················· | ···················· |

| | | | | | | | |
|---|---|---|---|---|---|---|---|
| Y | 5615 hCoV-19/Uganda/UG124/2020\|EPI_ISL_738031\|2020-09-0 | ···················· | ···················· | ····················· | ···················· | ···················· | ···················· |
| Y | 5616 hCoV-19/Uganda/UG125/2020\|EPI_ISL_738032\|2020-09-0 | ···················· | ···················· | ····················· | ···················· | ···················· | ···················· |
| Y | 5617 hCoV-19/Uganda/UG126/2020\|EPI_ISL_738033\|2020-09-0 | ···················· | ···················· | ····················· | ···················· | ···················· | ···················· |
| Y | 5618 hCoV-19/Uganda/UG127/2020\|EPI_ISL_738034\|2020-09-0 | ···················· | ···················· | ····················· | ···················· | ···················· | ···················· |
| Y | 5619 hCoV-19/Uganda/UG128/2020\|EPI_ISL_738035\|2020-09-0 | ···················· | ···················· | ····················· | ···················· | ···················· | ···················· |
| Y | 5620 hCoV-19/Uganda/UG129/2020\|EPI_ISL_738036\|2020-09-0 | ···················· | ···················· | ····················· | ···················· | ···················· | ···················· |
| Y | 5621 hCoV-19/Uganda/UG130/2020\|EPI_ISL_738037\|2020-09-0 | ···················· | ···················· | ····················· | ···················· | ···················· | ···················· |
| Y | 5622 hCoV-19/Uganda/UG131/2020\|EPI_ISL_738038\|2020-10-0 | ···················· | ···················· | ····················· | ···················· | ···················· | ···················· |
| Y | 5623 hCoV-19/Uganda/UG132/2020\|EPI_ISL_738039\|2020-10-0 | ···················· | ···················· | ····················· | ···················· | ···················· | ···················· |
| Y | 5624 hCoV-19/Uganda/UG133/2020\|EPI_ISL_738040\|2020-10-0 | ···················· | ···················· | ····················· | ···················· | ···················· | ···················· |
| Y | 5625 hCoV-19/Uganda/UG134/2020\|EPI_ISL_738041\|2020-10-0 | ···················· | ···················· | ····················· | ···················· | ···················· | ···················· |
| Y | 5626 hCoV-19/Uganda/UG135/2020\|EPI_ISL_738042\|2020-10-0 | ···················· | ···················· | ····················· | ···················· | ···················· | ···················· |
| Y | 5627 hCoV-19/Uganda/UG136/2020\|EPI_ISL_738043\|2020-10-0 | ···················· | ···················· | ····················· | ···················· | ···················· | ···················· |
| Y | 5628 hCoV-19/Italy/ABR-TE351205/2020\|EPI_ISL_738044\|202 | ···················· | ···················· | ····················· | ···················· | ···················· | ···················· |
| Y | 5629 hCoV-19/Taiwan/NTU36/2020\|EPI_ISL_738064\|2020-11-2 | ···················· | ···················· | ····················· | ···················· | ···················· | ···················· |
| Y | 5630 hCoV-19/Taiwan/NTU38/2020\|EPI_ISL_738065\|2020-11-2 | ···················· | ···················· | ····················· | ···················· | ···················· | ···················· |
| Y | 5631 hCoV-19/Switzerland/GE-621901/2020\|EPI_ISL_738085\| | ···················· | ···················· | ····················· | ···················· | ···················· | ···················· |
| Y | 5632 hCoV-19/Malaysia/HPUPM-ML0074/2020\|EPI_ISL_738086\| | ···················· | ···················· | ····················· | ···················· | ···················· | ···················· |
| Y | 5633 hCoV-19/Wales/PHWC-494313/2020\|EPI_ISL_726100\|2020 | ···················· | ···················· | ····················· | ···················· | ···················· | ···················· |
| Y | 5634 hCoV-19/Wales/PHWC-494322/2020\|EPI_ISL_726101\|2020 | ···················· | ···················· | ····················· | ···················· | ···················· | ···················· |
| Y | 5635 hCoV-19/Wales/PHWC-494331/2020\|EPI_ISL_726102\|2020 | ···················· | ···················· | ····················· | ···················· | ···················· | ···················· |
| Y | 5637 hCoV-19/Wales/PHWC-49436E/2020\|EPI_ISL_726104\|2020 | ···················· | ···················· | ····················· | ···················· | ···················· | ···················· |
| Y | 5638 hCoV-19/Wales/PHWC-49437D/2020\|EPI_ISL_726105\|2020 | ···················· | ···················· | ····················· | ···················· | ···················· | ···················· |
| Y | 5639 hCoV-19/Wales/PHWC-49438C/2020\|EPI_ISL_726106\|2020 | ···················· | ···················· | ····················· | ···················· | ···················· | ···················· |
| Y | 5640 hCoV-19/Wales/PHWC-49439B/2020\|EPI_ISL_726107\|2020 | ···················· | ···················· | ····················· | ···················· | ···················· | ···················· |
| Y | 5641 hCoV-19/Wales/PHWC-4943B9/2020\|EPI_ISL_726108\|2020 | ···················· | ···················· | ····················· | ···················· | ···················· | ···················· |
| Y | 5642 hCoV-19/Wales/PHWC-4943D7/2020\|EPI_ISL_726109\|2020 | ···················· | ···················· | ····················· | ···················· | ···················· | ···················· |
| Y | 5643 hCoV-19/Wales/PHWC-4943E6/2020\|EPI_ISL_726110\|2020 | ···················· | ···················· | ····················· | ···················· | ···················· | ···················· |
| Y | 5645 hCoV-19/Wales/PHWC-494410/2020\|EPI_ISL_726113\|2020 | ···················· | ···················· | ····················· | ···················· | ···················· | ···················· |
| Y | 5646 hCoV-19/Wales/PHWC-49443E/2020\|EPI_ISL_726115\|2020 | ···················· | ···················· | ····················· | ···················· | ···················· | ···················· |
| Y | 5647 hCoV-19/Wales/PHWC-49444D/2020\|EPI_ISL_726116\|2020 | ···················· | ···················· | ····················· | ···················· | ···················· | ···················· |
| Y | 5649 hCoV-19/Wales/PHWC-49446B/2020\|EPI_ISL_726118\|2020 | ···················· | ···················· | ····················· | ···················· | ···················· | ···················· |
| Y | 5650 hCoV-19/Wales/PHWC-49447A/2020\|EPI_ISL_726119\|2020 | ···················· | ···················· | ····················· | ···················· | ···················· | ···················· |
| Y | 5651 hCoV-19/Wales/PHWC-494489/2020\|EPI_ISL_726120\|2020 | ···················· | ···················· | ····················· | ···················· | ···················· | ···················· |
| Y | 5653 hCoV-19/Wales/PHWC-4944A7/2020\|EPI_ISL_726122\|2020 | ···················· | ···················· | ····················· | ···················· | ···················· | ···················· |
| Y | 5654 hCoV-19/Wales/PHWC-4944B6/2020\|EPI_ISL_726123\|2020 | ···················· | ···················· | ····················· | ···················· | ···················· | ···················· |
| Y | 5655 hCoV-19/Wales/PHWC-4944C5/2020\|EPI_ISL_726124\|2020 | ···················· | ···················· | ····················· | ···················· | ···················· | ···················· |
| Y | 5657 hCoV-19/Wales/PHWC-4944E3/2020\|EPI_ISL_726126\|2020 | ···················· | ···················· | ····················· | ···················· | ···················· | ···················· |
| Y | 5659 hCoV-19/Wales/PHWC-49450E/2020\|EPI_ISL_726128\|2020 | ···················· | ···················· | ····················· | ···················· | ···················· | ···················· |
| Y | 5660 hCoV-19/Wales/PHWC-49451D/2020\|EPI_ISL_726129\|2020 | ···················· | ···················· | ····················· | ···················· | ···················· | ···················· |
| Y | 5661 hCoV-19/Wales/PHWC-49454A/2020\|EPI_ISL_726131\|2020 | ···················· | ···················· | ····················· | ···················· | ···················· | ···················· |
| Y | 5662 hCoV-19/Wales/PHWC-494559/2020\|EPI_ISL_726132\|2020 | ···················· | ···················· | ····················· | ···················· | ···················· | ···················· |
| Y | 5663 hCoV-19/Wales/PHWC-494568/2020\|EPI_ISL_726133\|2020 | ···················· | ···················· | ····················· | ···················· | ···················· | ···················· |
| Y | 5664 hCoV-19/Wales/PHWC-494577/2020\|EPI_ISL_726134\|2020 | ···················· | ···················· | ····················· | ···················· | ···················· | ···················· |
| Y | 5665 hCoV-19/Wales/PHWC-494586/2020\|EPI_ISL_726135\|2020 | ···················· | ···················· | ····················· | ···················· | ···················· | ···················· |
| Y | 5667 hCoV-19/Wales/PHWC-4945A4/2020\|EPI_ISL_726137\|2020 | ···················· | ···················· | ····················· | ···················· | ···················· | ···················· |
| Y | 5669 hCoV-19/Wales/PHWC-4945D1/2020\|EPI_ISL_726140\|2020 | ···················· | ···················· | ····················· | ···················· | N··················· | ···················· |
| Y | 5670 hCoV-19/Wales/PHWC-4945E0/2020\|EPI_ISL_726141\|2020 | ···················· | ······A············· | ····················· | ···················· | ···················· | ···················· |
| Y | 5672 hCoV-19/Wales/PHWC-49460B/2020\|EPI_ISL_726143\|2020 | ···················· | ···················· | ····················· | ···················· | ···················· | ···················· |
| Y | 5674 hCoV-19/Wales/PHWC-494629/2020\|EPI_ISL_726145\|2020 | ···················· | ···················· | ····················· | ···················· | ···················· | ···················· |
| Y | 5675 hCoV-19/Wales/PHWC-494638/2020\|EPI_ISL_726146\|2020 | ···················· | ···················· | ····················· | ···················· | ···················· | ···················· |
| Y | 5676 hCoV-19/Wales/PHWC-494647/2020\|EPI_ISL_726147\|2020 | ···················· | ···················· | ····················· | ···················· | ···················· | ···················· |
| Y | 5677 hCoV-19/Wales/PHWC-494656/2020\|EPI_ISL_726148\|2020 | ···················· | ···················· | ····················· | ···················· | ···················· | ···················· |
| Y | 5678 hCoV-19/Wales/PHWC-494665/2020\|EPI_ISL_726149\|2020 | ···················· | ···················· | ····················· | ···················· | ···················· | ···················· |
| Y | 5679 hCoV-19/Wales/PHWC-494674/2020\|EPI_ISL_726150\|2020 | ···················· | ···················· | ····················· | ···················· | ···················· | ···················· |
| Y | 5680 hCoV-19/Wales/PHWC-494683/2020\|EPI_ISL_726151\|2020 | ···················· | ···················· | ····················· | ···················· | ···················· | ···················· |
| Y | 5681 hCoV-19/Wales/PHWC-494692/2020\|EPI_ISL_726152\|2020 | ···················· | ···················· | ····················· | ···················· | ···················· | ···················· |
| Y | 5683 hCoV-19/Wales/PHWC-4946B0/2020\|EPI_ISL_726154\|2020 | ···················· | ···················· | ····················· | ···················· | ···················· | ···················· |
| Y | 5684 hCoV-19/Wales/PHWC-4946CF/2020\|EPI_ISL_726155\|2020 | ···················· | ···················· | ····················· | ···················· | ···················· | ···················· |
| Y | 5685 hCoV-19/Wales/PHWC-4946DE/2020\|EPI_ISL_726156\|2020 | ···················· | ···················· | ····················· | ···················· | N··················· | ···················· |
| Y | 5686 hCoV-19/Wales/PHWC-4946ED/2020\|EPI_ISL_726157\|2020 | ···················· | ···················· | ····················· | ···················· | ···················· | ···················· |
| Y | 5687 hCoV-19/Wales/PHWC-4946FC/2020\|EPI_ISL_726158\|2020 | ···················· | ···················· | ····················· | ···················· | ···················· | ···················· |
| Y | 5688 hCoV-19/Wales/PHWC-494708/2020\|EPI_ISL_726159\|2020 | ···················· | ···················· | ····················· | ···················· | ···················· | ···················· |
| Y | 5689 hCoV-19/Wales/PHWC-494717/2020\|EPI_ISL_726160\|2020 | ···················· | ···················· | ····················· | ···················· | ···················· | ···················· |
| Y | 5690 hCoV-19/Wales/PHWC-494726/2020\|EPI_ISL_726161\|2020 | ···················· | ···················· | ····················· | ···················· | ···················· | ···················· |
| Y | 5691 hCoV-19/Wales/PHWC-494735/2020\|EPI_ISL_726162\|2020 | ···················· | ···················· | ····················· | ···················· | ···················· | ···················· |
| Y | 5692 hCoV-19/Wales/PHWC-494780/2020\|EPI_ISL_726163\|2020 | ···················· | ···················· | ····················· | ···················· | ···················· | ···················· |
| Y | 5693 hCoV-19/Wales/PHWC-49479F/2020\|EPI_ISL_726164\|2020 | ···················· | ···················· | ····················· | ···················· | ···················· | ···················· |
| Y | 5694 hCoV-19/Wales/PHWC-4947AE/2020\|EPI_ISL_726165\|2020 | ···················· | ···················· | ····················· | ···················· | ···················· | ···················· |
| Y | 5695 hCoV-19/Wales/PHWC-4947BD/2020\|EPI_ISL_726166\|2020 | ···················· | ···················· | ····················· | ···················· | ···················· | ···················· |
| Y | 5696 hCoV-19/Wales/PHWC-4947CC/2020\|EPI_ISL_726167\|2020 | ···················· | ···················· | ····················· | ···················· | ···················· | ···················· |
| Y | 5697 hCoV-19/Wales/PHWC-4947DB/2020\|EPI_ISL_726168\|2020 | ···················· | ···················· | ····················· | ···················· | ···················· | ···················· |
| Y | 5698 hCoV-19/Wales/PHWC-4947EA/2020\|EPI_ISL_726169\|2020 | ···················· | ···················· | ····················· | ···················· | ···················· | ···················· |
| Y | 5699 hCoV-19/Wales/PHWC-49493F/2020\|EPI_ISL_726190\|2020 | ···················· | ···················· | ····················· | ···················· | ···················· | ···················· |
| Y | 5701 hCoV-19/Wales/PHWC-49495D/2020\|EPI_ISL_726192\|2020 | ···················· | ···················· | ····················· | ···················· | ···················· | ···················· |
| Y | 5703 hCoV-19/Wales/PHWC-49497B/2020\|EPI_ISL_726194\|2020 | ···················· | ···················· | ····················· | ···················· | ···················· | ···················· |
| Y | 5704 hCoV-19/Wales/PHWC-494999/2020\|EPI_ISL_726196\|2020 | ···················· | ······A············· | ····················· | ···················· | ···················· | ···················· |
| Y | 5705 hCoV-19/Wales/PHWC-4949B7/2020\|EPI_ISL_726198\|2020 | ·T·················· | ···················· | ····················· | ···················· | ···················· | ······G············· |
| Y | 5706 hCoV-19/Wales/PHWC-4949C6/2020\|EPI_ISL_726199\|2020 | ···················· | ···················· | ····················· | ···················· | ···················· | ···················· |
| Y | 5708 hCoV-19/Wales/PHWC-499B07/2020\|EPI_ISL_727402\|2020 | ···················· | ···················· | ····················· | ···················· | ···················· | ···················· |

| | | | | | | | | | | |
|---|---|---|---|---|---|---|---|---|---|---|
| Y | 5712 | hCoV-19/Wales/PHWC-499B52/2020\|EPI_ISL_727406\|2020 | ··················· | ··················· | ···················· | ··················· | ··················· | ··················· | | |
| Y | 5713 | hCoV-19/Wales/PHWC-499B61/2020\|EPI_ISL_727407\|2020 | ··················· | ··················· | ···················· | ··················· | ··················· | ··················· | | |
| Y | 5716 | hCoV-19/Wales/PHWC-499BAD/2020\|EPI_ISL_727411\|2020 | ··················· | ··················· | ···················· | ··················· | ··················· | ··················· | | |
| Y | 5719 | hCoV-19/Wales/PHWC-499BF8/2020\|EPI_ISL_727416\|2020 | ··················· | ··················· | ···················· | ··················· | ··················· | ··················· | | |
| Y | 5720 | hCoV-19/Wales/PHWC-499C04/2020\|EPI_ISL_727417\|2020 | ··················· | ··················· | ···················· | ··················· | ··················· | ··················· | | |
| Y | 5722 | hCoV-19/Wales/PHWC-499C22/2020\|EPI_ISL_727419\|2020 | ··················· | ··················· | ···················· | ··················· | ··················· | ··················· | | |
| Y | 5723 | hCoV-19/Wales/PHWC-499C40/2020\|EPI_ISL_727421\|2020 | ··················· | ··················· | ···················· | ··················· | ··················· | ··················· | | |
| Y | 5724 | hCoV-19/Wales/PHWC-499C5F/2020\|EPI_ISL_727422\|2020 | ··················· | ··················· | ···················· | ··················· | ··················· | ··················· | | |
| Y | 5725 | hCoV-19/Wales/PHWC-499C6E/2020\|EPI_ISL_727423\|2020 | ··················· | ··················· | ···················· | ··················· | ··················· | ··················· | | |
| Y | 5728 | hCoV-19/Wales/PHWC-499CAA/2020\|EPI_ISL_727427\|2020 | ··················· | ··················· | ···················· | ··················· | ··················· | ··················· | | |
| Y | 5729 | hCoV-19/Wales/PHWC-499CB9/2020\|EPI_ISL_727428\|2020 | ··················· | ··················· | ···················· | ··················· | ··················· | ··················· | | |
| Y | 5730 | hCoV-19/Wales/PHWC-499CC8/2020\|EPI_ISL_727429\|2020 | ··················· | ··················· | ···················· | ··················· | ··················· | ··················· | | |
| Y | 5733 | hCoV-19/Wales/PHWC-499D10/2020\|EPI_ISL_727434\|2020 | ··················· | ··················· | ···················· | ··················· | ··················· | ··················· | | |
| Y | 5734 | hCoV-19/Wales/PHWC-499D4D/2020\|EPI_ISL_727437\|2020 | ··················· | ··················· | ···················· | ··················· | N·················· | ··················· | | |
| Y | 5736 | hCoV-19/Wales/PHWC-499D7A/2020\|EPI_ISL_727440\|2020 | ··················· | ··················· | ···················· | ··················· | ··················· | ··················· | | |
| Y | 5737 | hCoV-19/Wales/PHWC-499D89/2020\|EPI_ISL_727441\|2020 | ··················· | ··················· | ···················· | ··················· | ··················· | ··················· | | |
| Y | 5738 | hCoV-19/Wales/PHWC-499D98/2020\|EPI_ISL_727442\|2020 | ··················· | ··················· | ···················· | ··················· | ··················· | ··················· | | |
| Y | 5740 | hCoV-19/Wales/PHWC-499DC5/2020\|EPI_ISL_727445\|2020 | ··················· | ··················· | ···················· | ··················· | ··················· | ··················· | | |
| Y | 5741 | hCoV-19/Wales/PHWC-499DD4/2020\|EPI_ISL_727446\|2020 | ··················· | ··················· | ···················· | ··················· | ··················· | ··················· | | |
| Y | 5742 | hCoV-19/Wales/PHWC-499DE3/2020\|EPI_ISL_727447\|2020 | ··················· | ··················· | ···················· | ··················· | ··················· | ··················· | | |
| Y | 5743 | hCoV-19/Wales/PHWC-499DF2/2020\|EPI_ISL_727448\|2020 | ··················· | ··················· | ···················· | ··················· | ··················· | ··················· | | |
| Y | 5747 | hCoV-19/Wales/PHWC-499E4A/2020\|EPI_ISL_727453\|2020 | ··················· | ··················· | ···················· | ··················· | ··················· | ··················N | 1 | |
| Y | 5748 | hCoV-19/Wales/PHWC-499E68/2020\|EPI_ISL_727455\|2020 | ··················· | ··················· | ···················· | ··················· | ··················· | ··················· | | |
| Y | 5749 | hCoV-19/Wales/PHWC-499E77/2020\|EPI_ISL_727456\|2020 | ·T················· | ··················· | ···················· | ··················· | ··················· | ·····G············· | | |
| Y | 5750 | hCoV-19/Wales/PHWC-499E86/2020\|EPI_ISL_727457\|2020 | ··················· | ··················· | ···················· | ··················· | ··················· | ··················· | | |
| Y | 5753 | hCoV-19/Wales/PHWC-499EC2/2020\|EPI_ISL_727461\|2020 | ··················· | ··················· | ···················· | ··················· | ··················· | ··················· | | |
| Y | 5757 | hCoV-19/Wales/PHWC-499F0B/2020\|EPI_ISL_727465\|2020 | ··················· | ··················· | ···················· | ··················· | ··················· | ··················· | | |
| Y | 5758 | hCoV-19/Wales/PHWC-499F1A/2020\|EPI_ISL_727466\|2020 | ··················· | ··················· | ···················· | ··················· | ··················· | ··················· | | |
| Y | 5759 | hCoV-19/Wales/PHWC-499F29/2020\|EPI_ISL_727467\|2020 | ··················· | ··················· | ···················· | ··················· | ··················· | ··················· | | |
| Y | 5763 | hCoV-19/Wales/PHWC-499F92/2020\|EPI_ISL_727474\|2020 | ··················· | ··················· | ···················· | ··················· | ··················· | ··················· | | |
| Y | 5765 | hCoV-19/Wales/PHWC-499FCF/2020\|EPI_ISL_727477\|2020 | ··················· | ··················· | ···················· | ··················· | ··················· | ··················· | | |
| Y | 5768 | hCoV-19/Wales/PHWC-499FFC/2020\|EPI_ISL_727480\|2020 | ··················· | ··················· | ···················· | ··················· | ··················· | ··················· | | |
| Y | 5769 | hCoV-19/Wales/PHWC-49A007/2020\|EPI_ISL_727481\|2020 | ··················· | ··················· | ···················· | ··················· | ··················· | ··················· | | |
| Y | 5771 | hCoV-19/Wales/PHWC-49A025/2020\|EPI_ISL_727483\|2020 | ··················· | ··················· | ···················· | ··················· | ··················· | ··················· | | |
| Y | 5772 | hCoV-19/Wales/PHWC-49A061/2020\|EPI_ISL_727486\|2020 | ··················· | ··················· | ···················· | ··················· | ··················· | ··················· | | |
| Y | 5773 | hCoV-19/Wales/PHWC-49A070/2020\|EPI_ISL_727487\|2020 | ··················· | ··················· | ···················· | ··················· | ··················· | ··················· | | |
| Y | 5774 | hCoV-19/Wales/PHWC-49A09E/2020\|EPI_ISL_727489\|2020 | ··················· | ··················· | ···················· | ··················· | ··················· | ··················· | | |
| Y | 5775 | hCoV-19/Wales/PHWC-49A0AD/2020\|EPI_ISL_727490\|2020 | ··················· | ··················· | ···················· | ··················· | ··················· | ··················· | | |
| Y | 5776 | hCoV-19/Wales/PHWC-49A0BC/2020\|EPI_ISL_727491\|2020 | ··················· | ··················· | ···················· | ··················· | NNN················ | ··················· | | 1 |
| Y | 5777 | hCoV-19/Wales/PHWC-49A0CB/2020\|EPI_ISL_727492\|2020 | ··················· | ··················· | ···················· | ··················· | ··················· | ··················· | | |
| Y | 5778 | hCoV-19/Wales/PHWC-49A0E9/2020\|EPI_ISL_727494\|2020 | ··················· | ··················· | ···················· | ··················· | NNN················ | ··················· | | 1 |
| Y | 5779 | hCoV-19/Wales/PHWC-49A0F8/2020\|EPI_ISL_727495\|2020 | ··················· | ··················· | ···················· | ··················· | ··················· | ··················· | | |
| Y | 5780 | hCoV-19/Wales/PHWC-49A104/2020\|EPI_ISL_727496\|2020 | ··················· | ··················· | ···················· | ··················· | ··················· | ··················· | | |
| Y | 5781 | hCoV-19/Wales/PHWC-49A113/2020\|EPI_ISL_727497\|2020 | ··················· | ··················· | ···················· | ··················· | ··················· | ··················· | | |
| Y | 5782 | hCoV-19/Wales/PHWC-49A122/2020\|EPI_ISL_727498\|2020 | ··················· | ··················· | ···················· | ··················· | ··················· | ··················· | | |
| Y | 5783 | hCoV-19/Wales/PHWC-49A131/2020\|EPI_ISL_727499\|2020 | ··················· | ··················· | ···················· | ··················· | ··················· | ··················· | | |
| Y | 5784 | hCoV-19/Netherlands/OV-RIVM-20493/2020\|EPI_ISL_728 | ··················· | ··················· | ···················· | ··················· | ··················· | ··················· | | |
| Y | 5785 | hCoV-19/Netherlands/OV-RIVM-20494/2020\|EPI_ISL_728 | ··················· | ··················· | ···················· | ··················· | ··················· | ··················· | | |
| Y | 5786 | hCoV-19/Netherlands/OV-RIVM-20495/2020\|EPI_ISL_728 | ··················· | ··················· | ···················· | ··················· | ··················· | ··················· | | |
| Y | 5787 | hCoV-19/Netherlands/OV-RIVM-20496/2020\|EPI_ISL_728 | ··················· | ··················· | ···················· | ··················· | ··················· | ··················· | | |
| Y | 5788 | hCoV-19/Netherlands/OV-RIVM-20497/2020\|EPI_ISL_728 | ··················· | ··················· | ···················· | ··················· | ··················· | ··················· | | |
| Y | 5789 | hCoV-19/Netherlands/OV-RIVM-20498/2020\|EPI_ISL_728 | ··················· | ··················· | ···················· | ··················· | ··················· | ··················· | | |
| Y | 5790 | hCoV-19/Netherlands/OV-RIVM-20499/2020\|EPI_ISL_728 | ··················· | ··················· | ···················· | ··················· | ··················· | ··················· | | |
| Y | 5791 | hCoV-19/Netherlands/OV-RIVM-20500/2020\|EPI_ISL_728 | ··················· | ··················· | ···················· | ··················· | ··················· | ··················· | | |
| Y | 5792 | hCoV-19/Netherlands/OV-RIVM-20501/2020\|EPI_ISL_728 | ··················· | ··················· | ···················· | ··················· | ··················· | ··················· | | |
| Y | 5793 | hCoV-19/Netherlands/OV-RIVM-20502/2020\|EPI_ISL_728 | ··················· | ··················· | ···················· | ··················· | ··················· | ··················· | | |
| Y | 5794 | hCoV-19/Netherlands/OV-RIVM-20503/2020\|EPI_ISL_728 | ··················· | ··················· | ···················· | ··················· | ··················· | ··················· | | |
| Y | 5795 | hCoV-19/Netherlands/OV-RIVM-20504/2020\|EPI_ISL_728 | ··················· | ··················· | ···················· | ··················· | ··················· | ··················· | | |
| Y | 5796 | hCoV-19/Netherlands/OV-RIVM-20505/2020\|EPI_ISL_728 | ··················· | ··················· | ···················· | ··················· | ··················· | ··················· | | |
| Y | 5797 | hCoV-19/Netherlands/LI-RIVM-20506/2020\|EPI_ISL_728 | ··················· | ··················· | ···················· | ··················· | ··················· | ··················· | | |
| Y | 5798 | hCoV-19/Netherlands/LI-RIVM-20507/2020\|EPI_ISL_728 | ··················· | ··················· | ···················· | ··················· | ··················· | ··················· | | |
| Y | 5799 | hCoV-19/Netherlands/OV-RIVM-20508/2020\|EPI_ISL_728 | ··················· | ··················· | ···················· | ··················· | ··················· | ··················· | | |
| Y | 5800 | hCoV-19/Netherlands/OV-RIVM-20509/2020\|EPI_ISL_728 | ··················· | ··················· | ···················· | ··················· | ··················· | ··················· | | |
| Y | 5801 | hCoV-19/Netherlands/OV-RIVM-20510/2020\|EPI_ISL_728 | ··················· | ··················· | ···················· | ··················· | ··················· | ··················· | | |
| Y | 5802 | hCoV-19/Netherlands/OV-RIVM-20511/2020\|EPI_ISL_728 | ··················· | ··················· | ···················· | ··················· | ··················· | ··················· | | |
| Y | 5803 | hCoV-19/Netherlands/OV-RIVM-20512/2020\|EPI_ISL_728 | ··················· | ··················· | ···················· | ··················· | ··················· | ··················· | | |
| Y | 5804 | hCoV-19/Netherlands/OV-RIVM-20513/2020\|EPI_ISL_728 | ··················· | ··················· | ···················· | ··················· | ··················· | ··················· | | |
| Y | 5805 | hCoV-19/Netherlands/OV-RIVM-20514/2020\|EPI_ISL_728 | ··················· | ··················· | ···················· | ··················· | ··················· | ··················· | | |
| Y | 5806 | hCoV-19/Netherlands/OV-RIVM-20515/2020\|EPI_ISL_728 | ··················· | ··················· | ···················· | ··················· | ··················· | ··················· | | |
| Y | 5807 | hCoV-19/Netherlands/OV-RIVM-20516/2020\|EPI_ISL_728 | ··················· | ··················· | ···················· | ··················· | ··················· | ··················· | | |
| Y | 5808 | hCoV-19/Netherlands/OV-RIVM-20517/2020\|EPI_ISL_728 | ··················· | ··················· | ···················· | ··················· | ·······G··········· | ··················· | | |
| Y | 5809 | hCoV-19/Netherlands/OV-RIVM-20518/2020\|EPI_ISL_728 | ··················· | ··················· | ···················· | ··················· | ··················· | ··················· | | |
| Y | 5810 | hCoV-19/Netherlands/GE-RIVM-20520/2020\|EPI_ISL_728 | ··················· | ··················· | ···················· | ··················· | ··················· | ··················· | | |
| Y | 5811 | hCoV-19/Netherlands/GE-RIVM-20521/2020\|EPI_ISL_728 | ··················· | ··················· | ···················· | ··················· | ··················· | ··················· | | |
| Y | 5812 | hCoV-19/Netherlands/GE-RIVM-20522/2020\|EPI_ISL_728 | ··················· | ··················· | ···················· | ··················· | ··················· | ··················· | | |
| Y | 5813 | hCoV-19/Netherlands/GE-RIVM-20523/2020\|EPI_ISL_728 | ··················· | ··················· | ···················· | ··················· | ··················· | ··················· | | |
| Y | 5814 | hCoV-19/Netherlands/GE-RIVM-20524/2020\|EPI_ISL_728 | ··················· | ··················· | ···················· | ··················· | ··················· | ··················· | | |
| Y | 5815 | hCoV-19/Netherlands/GE-RIVM-20525/2020\|EPI_ISL_728 | ··················· | ··················· | ···················· | ··················· | ··················· | ··················· | | |
| Y | 5816 | hCoV-19/Netherlands/GE-RIVM-20526/2020\|EPI_ISL_728 | ··················· | ··················· | ···················· | ··················· | ··················· | ··················· | | |
| Y | 5817 | hCoV-19/Netherlands/GE-RIVM-20527/2020\|EPI_ISL_728 | ··················· | ··················· | ···················· | ··················· | ··················· | ··················· | | |

| | | | | | | | | |
|---|---|---|---|---|---|---|---|---|
| Y | 5818 | hCoV-19/Netherlands/GE-RIVM-20528/2020\|EPI_ISL_728 | ···················· | ···················· | ···················· | ···················· | ···················· | ···················· |
| Y | 5819 | hCoV-19/Netherlands/GE-RIVM-20529/2020\|EPI_ISL_728 | ···················· | ···················· | ···················· | ···················· | ···················· | ···················· |
| Y | 5820 | hCoV-19/Netherlands/NB-RIVM-20530/2020\|EPI_ISL_728 | ···················· | ···················· | ···················· | ···················· | ···················· | ···················· |
| Y | 5821 | hCoV-19/Netherlands/NH-RIVM-20531/2020\|EPI_ISL_728 | ···················· | ···················· | ···················· | ···················· | ···················· | ···················· |
| Y | 5822 | hCoV-19/Netherlands/ZH-RIVM-20532/2020\|EPI_ISL_728 | ···················· | ···················· | ···················· | ···················· | ······T············· | ···················· |
| Y | 5823 | hCoV-19/Netherlands/OV-RIVM-20533/2020\|EPI_ISL_728 | ···················· | ···················· | ···················· | ···················· | ···················· | ···················· |
| Y | 5824 | hCoV-19/Netherlands/OV-RIVM-20534/2020\|EPI_ISL_728 | ···················· | ···················· | ···················· | ···················· | ···················· | ···················· |
| Y | 5825 | hCoV-19/Netherlands/OV-RIVM-20535/2020\|EPI_ISL_728 | ···················· | ···················· | ···················· | ···················· | ···················· | ···················· |
| Y | 5826 | hCoV-19/Netherlands/OV-RIVM-20536/2020\|EPI_ISL_728 | ···················· | ···················· | ···················· | ···················· | ···················· | ···················· |
| Y | 5827 | hCoV-19/Netherlands/OV-RIVM-20537/2020\|EPI_ISL_728 | ···················· | ···················· | ···················· | ···················· | ···················· | ···················· |
| Y | 5828 | hCoV-19/Netherlands/OV-RIVM-20538/2020\|EPI_ISL_728 | ···················· | ···················· | ···················· | ···················· | ···················· | ···················· |
| Y | 5829 | hCoV-19/Netherlands/OV-RIVM-20539/2020\|EPI_ISL_728 | ···················· | ···················· | ···················· | ···················· | ···················· | ···················· |
| Y | 5830 | hCoV-19/Netherlands/OV-RIVM-20540/2020\|EPI_ISL_728 | ···················· | ···················· | ···················· | ···················· | ···················· | ···················· |
| Y | 5831 | hCoV-19/Netherlands/OV-RIVM-20541/2020\|EPI_ISL_728 | ···················· | ···················· | ···················· | ···················· | ···················· | ···················· |
| Y | 5832 | hCoV-19/Netherlands/GE-RIVM-20542/2020\|EPI_ISL_728 | ···················· | ···················· | ···················· | ···················· | ···················· | ···················· |
| Y | 5833 | hCoV-19/Netherlands/GE-RIVM-20543/2020\|EPI_ISL_728 | ···················· | ···················· | ···················· | ···················· | ···················· | ···················· |
| Y | 5834 | hCoV-19/Netherlands/GE-RIVM-20544/2020\|EPI_ISL_728 | ···················· | ·A·················· | ···················· | ···················· | ···················· | ···A················ |
| Y | 5835 | hCoV-19/Netherlands/GE-RIVM-20545/2020\|EPI_ISL_728 | ···················· | ···················· | ···················· | ···················· | ···················· | ···················· |
| Y | 5836 | hCoV-19/Netherlands/GE-RIVM-20546/2020\|EPI_ISL_728 | ···T················ | ···················· | ···················· | ···················· | ···················· | ···················· |
| Y | 5837 | hCoV-19/Netherlands/GE-RIVM-20547/2020\|EPI_ISL_728 | ···················· | ···················· | ···················· | ···················· | ···················· | ···················· |
| Y | 5838 | hCoV-19/Netherlands/GE-RIVM-20548/2020\|EPI_ISL_728 | ···················· | ···················· | ···················· | ···················· | ···················· | ···················· |
| Y | 5839 | hCoV-19/Belgium/rega-0416498/2020\|EPI_ISL_734590\|2 | ···················· | ···················· | ···················· | ···················· | ···················· | ···················· |
| Y | 5840 | hCoV-19/Belgium/rega-0416499/2020\|EPI_ISL_734591\|2 | ···················· | ···················· | ···················· | ···················· | ···················· | ···················· |
| Y | 5841 | hCoV-19/England/ALDP-C5851D/2020\|EPI_ISL_735900\|20 | ···················· | ···················· | ···················· | ···················· | ···················· | ···A················ |
| Y | 5842 | hCoV-19/England/ALDP-C58647/2020\|EPI_ISL_735901\|20 | ···················· | ···················· | ···················· | ···················· | ···················· | ···················· |
| Y | 5843 | hCoV-19/England/ALDP-C585FF/2020\|EPI_ISL_735902\|20 | ···················· | ···················· | ···················· | ···················· | ···················· | ···A················ |
| Y | 5844 | hCoV-19/England/ALDP-C57FC5/2020\|EPI_ISL_735904\|20 | ···················· | ···················· | ···················· | ···················· | ···················· | ···················· |
| Y | 5845 | hCoV-19/England/ALDP-C58D06/2020\|EPI_ISL_735905\|20 | ···················· | ···················· | ···················· | ···················· | ···················· | ···················· |
| Y | 5846 | hCoV-19/England/ALDP-C58568/2020\|EPI_ISL_735907\|20 | ···················· | ···················· | ···················· | ···················· | ···················· | ···················· |
| Y | 5847 | hCoV-19/England/ALDP-C57C37/2020\|EPI_ISL_735908\|20 | ···················· | ···················· | ···················· | ···················· | ···················· | ···················· |
| Y | 5848 | hCoV-19/England/QEUH-C5BCF7/2020\|EPI_ISL_735909\|20 | ···················· | ·········C·········· | ···················· | ···················· | ···················· | ···················· |
| Y | 5849 | hCoV-19/England/ALDP-C58173/2020\|EPI_ISL_735910\|20 | ···················· | ···················· | ···················· | ···················· | ···················· | ···················· |
| Y | 5850 | hCoV-19/England/ALDP-C57F10/2020\|EPI_ISL_735911\|20 | ···················· | ···················· | ···················· | ···················· | ···················· | ···················· |
| Y | 5851 | hCoV-19/Portugal/PT2109/2020\|EPI_ISL_738112\|2020-1 | ···················· | ···················· | ···················· | ···················· | ···················· | ···················· |
| Y | 5852 | hCoV-19/Portugal/PT2111/2020\|EPI_ISL_738113\|2020-1 | ···················· | ···················· | ···················· | ···················· | ···················· | ···················· |
| Y | 5853 | hCoV-19/Portugal/PT2113/2020\|EPI_ISL_738114\|2020-1 | ···················· | ···················· | ···················· | ···················· | ···················· | ···················· |
| Y | 5854 | hCoV-19/Portugal/PT2081/2020\|EPI_ISL_738115\|2020-1 | ···················· | ···················· | ···················· | ···················· | ···················· | ···················· |
| Y | 5855 | hCoV-19/Portugal/PT2082/2020\|EPI_ISL_738116\|2020-1 | ···················· | ···················· | ···················· | ···················· | ···················· | ···················· |
| Y | 5856 | hCoV-19/Portugal/PT2085/2020\|EPI_ISL_738117\|2020-1 | ···················· | ···················· | ···················· | ···················· | ···················· | ···················· |
| Y | 5857 | hCoV-19/Portugal/PT2086/2020\|EPI_ISL_738118\|2020-1 | ···················· | ···················· | ···················· | ···················· | ···················· | ···················· |
| Y | 5858 | hCoV-19/Portugal/PT2087/2020\|EPI_ISL_738119\|2020-1 | ···················· | ···················· | ···················· | ···················· | ···················· | ···················· |
| Y | 5859 | hCoV-19/Portugal/PT2090/2020\|EPI_ISL_738120\|2020-1 | ···················· | ···················· | ···················· | ···················· | ···················· | ···················· |
| Y | 5860 | hCoV-19/Italy/CAM-IZSM-87503-NIKY/2020\|EPI_ISL_738 | ···················· | ···················· | ···················· | ···················· | ······T············· | ···················· |
| Y | 5861 | hCoV-19/Italy/CAM-IZSM-115255/2020\|EPI_ISL_738122\| | ···················· | ···················· | ···················· | ···················· | ······T····G········ | ···················· |
| Y | 5862 | hCoV-19/Italy/CAM-IZSM-115372/2020\|EPI_ISL_738124\| | ···················· | ···················· | ···················· | ···················· | ······T····G········ | ···················· |
| Y | 5863 | hCoV-19/Italy/CAM-IZSM-118588/2020\|EPI_ISL_738125\| | ···················· | ···················· | ···················· | ···················· | ···················· | ···················· |
| Y | 5864 | hCoV-19/Italy/CAM-IZSM-119242/2020\|EPI_ISL_738126\| | ···················· | ···················· | ···················· | ···················· | ···················· | ···················· |
| Y | 5865 | hCoV-19/Italy/CAM-IZSM-119627/2020\|EPI_ISL_738127\| | ···················· | ···················· | ···················· | ···················· | ···················· | ···················· |
| Y | 5866 | hCoV-19/Italy/CAM-IZSM-119235/2020\|EPI_ISL_738128\| | ···················· | ···················· | ···················· | ···················· | ···················· | ···················· |
| Y | 5867 | hCoV-19/Italy/CAM-IZSM-88763-ORS/2020\|EPI_ISL_7381 | ···················· | ···················· | ···················· | ···················· | ···················· | ···················· |
| Y | 5868 | hCoV-19/Italy/CAM-IZSM-118376/2020\|EPI_ISL_738130\| | ···················· | ···················· | ···················· | ···················· | ······T············· | ···················· |
| Y | 5869 | hCoV-19/Italy/CAM-IZSM-118611/2020\|EPI_ISL_738131\| | ···················· | ···················· | ···················· | ···················· | ···················· | ···················· |
| Y | 5870 | hCoV-19/Italy/CAM-IZSM-124821/2020\|EPI_ISL_738132\| | ···················· | ···················· | ···················· | ···················· | ······T············· | ···················· |
| Y | 5871 | hCoV-19/Australia/NSW3724/2020\|EPI_ISL_738137\|2020 | ···················· | ···················· | ···················· | ···················· | ···················· | ···················· |
| Y | 5872 | hCoV-19/Italy/APU-POLBA04/2020\|EPI_ISL_738144\|2020 | ···················· | ···················· | ···················· | ···················· | ······T············· | ···················· |
| Y | 5873 | hCoV-19/Italy/TUS-C44/2020\|EPI_ISL_738147\|2020-02- | ···················· | ···················· | ···················· | ···················· | ···················· | ···················· |
| Y | 5874 | hCoV-19/USA/TX-DSHS-1797/2020\|EPI_ISL_738170\|2020- | ···················· | ····T··············· | ···················· | ···················· | ···················· | ···················· |
| Y | 5875 | hCoV-19/USA/TX-DSHS-1798/2020\|EPI_ISL_738171\|2020- | ···················· | ·············A······ | ···················· | ···················· | ···················· | ···················· |
| Y | 5876 | hCoV-19/USA/TX-DSHS-1799/2020\|EPI_ISL_738172\|2020- | ···················· | ···················· | ···················· | ···················· | ···················· | ···················· |
| Y | 5877 | hCoV-19/USA/TX-DSHS-1802/2020\|EPI_ISL_738173\|2020- | ···················· | ···················· | ···················· | ···················· | ···················· | ···················· |
| Y | 5878 | hCoV-19/USA/TX-DSHS-1809/2020\|EPI_ISL_738179\|2020- | ···················· | ···················· | ···················· | ···················· | ···················· | ···················· |
| Y | 5879 | hCoV-19/USA/TX-DSHS-1810/2020\|EPI_ISL_738180\|2020- | ···················· | ···················· | ···················· | ···················· | ···················· | ···················· |
| Y | 5880 | hCoV-19/USA/TX-DSHS-1811/2020\|EPI_ISL_738181\|2020- | ···················· | ···················· | ···················· | ···················· | ···················· | ···················· |
| Y | 5881 | hCoV-19/USA/TX-DSHS-1814/2020\|EPI_ISL_738182\|2020- | ···················· | ···················· | ···················· | ···················· | ···················· | ···················· |
| Y | 5882 | hCoV-19/USA/TX-DSHS-1816/2020\|EPI_ISL_738183\|2020- | ···················· | ···················· | ···················· | ···················· | ···················· | ···················· |
| Y | 5883 | hCoV-19/USA/TX-DSHS-1817/2020\|EPI_ISL_738184\|2020- | ···················· | ·A·················· | ···················· | ···················· | ···················· | ···················· |
| Y | 5884 | hCoV-19/USA/TX-DSHS-1818/2020\|EPI_ISL_738185\|2020- | ···················· | ···················· | ···················· | ···················· | ···················· | ···················· |
| Y | 5885 | hCoV-19/USA/TX-DSHS-1819/2020\|EPI_ISL_738186\|2020- | ···················· | ···················· | ···················· | ···················· | ···················· | ···················· |
| Y | 5886 | hCoV-19/USA/TX-DSHS-1820/2020\|EPI_ISL_738187\|2020- | ···················· | ···················· | ···················· | ···················· | ···················· | ···················· |
| Y | 5887 | hCoV-19/USA/TX-DSHS-1821/2020\|EPI_ISL_738188\|2020- | ···················· | ···················· | ···················· | ···················· | ···················· | ···················· |
| Y | 5888 | hCoV-19/USA/TX-DSHS-1822/2020\|EPI_ISL_738189\|2020- | ···················· | ···················· | ···················· | ···················· | ···················· | ···················· |
| Y | 5889 | hCoV-19/Wales/PHWC-4947F9/2020\|EPI_ISL_726170\|2020 | ···················· | ···················· | ···················· | ···················· | ···················· | ···················· |
| Y | 5890 | hCoV-19/Wales/PHWC-494805/2020\|EPI_ISL_726171\|2020 | ···················· | ···················· | ···················· | ···················· | ···················· | ···················· |
| Y | 5891 | hCoV-19/Wales/PHWC-494814/2020\|EPI_ISL_726172\|2020 | ···················· | ···················· | ···················· | ···················· | ···················· | ···················· |
| Y | 5892 | hCoV-19/Wales/PHWC-494823/2020\|EPI_ISL_726173\|2020 | ···················· | ···················· | ···················· | ···················· | ···················· | ···················· |
| Y | 5893 | hCoV-19/Wales/PHWC-494832/2020\|EPI_ISL_726174\|2020 | ···················· | ···················· | ···················· | ···················· | ···················· | ···················· |
| Y | 5894 | hCoV-19/Wales/PHWC-494841/2020\|EPI_ISL_726175\|2020 | ···················· | ······A············· | ···················· | ···················· | ···················· | ···················· |
| Y | 5895 | hCoV-19/Wales/PHWC-49486F/2020\|EPI_ISL_726177\|2020 | ···················· | ···················· | ···················· | ···················· | ···················· | ···················· |
| Y | 5896 | hCoV-19/Wales/PHWC-49487E/2020\|EPI_ISL_726178\|2020 | ···················· | ···················· | ···················· | ···················· | ···················· | ···················· |
| Y | 5897 | hCoV-19/Wales/PHWC-49488D/2020\|EPI_ISL_726179\|2020 | ···················· | ···················· | ···················· | ···················· | ···················· | ···················· |

| | | | | | | | | |
|---|---|---|---|---|---|---|---|---|
| Y | 5898 | hCoV-19/Wales/PHWC-49489C/2020\|EPI_ISL_726180\|2020 | ···················· | ···················· | ····················· | ···················· | ···················· | ···················· |
| Y | 5899 | hCoV-19/Wales/PHWC-4948BA/2020\|EPI_ISL_726182\|2020 | ···················· | ···················· | ····················· | ···················· | ···················· | ···················· |
| Y | 5900 | hCoV-19/Wales/PHWC-4948C9/2020\|EPI_ISL_726183\|2020 | ···················· | ···················· | ····················· | ···················· | ···················· | ···················· |
| Y | 5901 | hCoV-19/Wales/PHWC-4948E7/2020\|EPI_ISL_726185\|2020 | ···················· | ···················· | ····················· | ···················· | ···················· | ···················· |
| Y | 5902 | hCoV-19/Wales/PHWC-4948F6/2020\|EPI_ISL_726186\|2020 | ···················· | ···················· | ····················· | ···················· | ···················· | ···················· |
| Y | 5903 | hCoV-19/Wales/PHWC-494902/2020\|EPI_ISL_726187\|2020 | ···················· | ···················· | ····················· | ···················· | ···················· | ···················· |
| Y | 5904 | hCoV-19/Wales/PHWC-494911/2020\|EPI_ISL_726188\|2020 | ···················· | ···················· | ····················· | ···················· | ···················· | ···················· |
| Y | 5905 | hCoV-19/Wales/PHWC-494920/2020\|EPI_ISL_726189\|2020 | ···················· | ···················· | ····················· | ···················· | ···················· | ···················· |
| Y | 5906 | hCoV-19/Portugal/PT1769/2020\|EPI_ISL_732000\|2020-1 | ···················· | ···················· | ····················· | ···················· | ···················· | ···················· |
| Y | 5907 | hCoV-19/Portugal/PT1770/2020\|EPI_ISL_732001\|2020-1 | ···················· | ···················· | ····················· | ···················· | ···················· | ···················· |
| Y | 5908 | hCoV-19/Portugal/PT1771/2020\|EPI_ISL_732002\|2020-1 | ···················· | ···················· | ····················· | ···················· | ···················· | ···················· |
| Y | 5909 | hCoV-19/Portugal/PT1772/2020\|EPI_ISL_732003\|2020-1 | ···················· | ···················· | ····················· | ···················· | ···················· | ···················· |
| Y | 5910 | hCoV-19/Portugal/PT1773/2020\|EPI_ISL_732004\|2020-1 | ···················· | ···················· | ····················· | ···················· | ···················· | ···················· |
| Y | 5911 | hCoV-19/Portugal/PT1774/2020\|EPI_ISL_732005\|2020-1 | ···················· | ···················· | ····················· | ···················· | ···················· | ···················· |
| Y | 5912 | hCoV-19/Portugal/PT1775/2020\|EPI_ISL_732006\|2020-1 | ···················· | ···················· | ····················· | ···················· | ···················· | ···················· |
| Y | 5913 | hCoV-19/Portugal/PT1776/2020\|EPI_ISL_732007\|2020-1 | ···················· | ···················· | ····················· | ···················· | ···················· | ···················· |
| Y | 5914 | hCoV-19/Portugal/PT1779/2020\|EPI_ISL_732008\|2020-1 | ···················· | ···················· | ····················· | ···················· | ···················· | ···················· |
| Y | 5915 | hCoV-19/Portugal/PT1780/2020\|EPI_ISL_732009\|2020-1 | ···················· | ···················· | ····················· | ···················· | ···················· | ···················· |
| Y | 5916 | hCoV-19/Portugal/PT1785/2020\|EPI_ISL_732014\|2020-1 | ···················· | ···················· | ····················· | ···················· | ···················· | ···················· |
| Y | 5917 | hCoV-19/Portugal/PT1791/2020\|EPI_ISL_732019\|2020-1 | ···················· | ···················· | ····················· | ···················· | ···················· | ···················· |
| Y | 5918 | hCoV-19/Portugal/PT1792/2020\|EPI_ISL_732020\|2020-1 | ···················· | ···················· | ····················· | ···················· | ···················· | ···················· |
| Y | 5919 | hCoV-19/Portugal/PT1793/2020\|EPI_ISL_732021\|2020-1 | ···················· | ···················· | ····················· | ···················· | ···················· | ···················· |
| Y | 5920 | hCoV-19/Portugal/PT1795/2020\|EPI_ISL_732022\|2020-1 | ···················· | ···················· | ····················· | ···················· | ···················· | ···················· |
| Y | 5921 | hCoV-19/Portugal/PT1796/2020\|EPI_ISL_732023\|2020-1 | ···················· | ···················· | ····················· | ···················· | ···················· | ···················· |
| Y | 5922 | hCoV-19/Portugal/PT1797/2020\|EPI_ISL_732024\|2020-1 | ···················· | ···················· | ····················· | ···················· | ···················· | ···················· |
| Y | 5923 | hCoV-19/Portugal/PT1798/2020\|EPI_ISL_732025\|2020-1 | ···················· | ···················· | ····················· | ···················· | ···················· | ···················· |
| Y | 5924 | hCoV-19/Portugal/PT1800/2020\|EPI_ISL_732026\|2020-1 | ···················· | ···················· | ····················· | ···················· | ···················· | ···················· |
| Y | 5925 | hCoV-19/Portugal/PT1801/2020\|EPI_ISL_732027\|2020-1 | ···················· | ···················· | ····················· | ···················· | ···················· | ···················· |
| Y | 5926 | hCoV-19/Portugal/PT1803/2020\|EPI_ISL_732028\|2020-1 | ···················· | ···················· | ····················· | ···················· | ···················· | ···················· |
| Y | 5927 | hCoV-19/Portugal/PT1804/2020\|EPI_ISL_732029\|2020-1 | ···················· | ···················· | ····················· | ···················· | ···················· | ···················· |
| Y | 5928 | hCoV-19/Portugal/PT1805/2020\|EPI_ISL_732030\|2020-1 | ···················· | ···················· | ····················· | ···················· | ···················· | ···················· |
| Y | 5929 | hCoV-19/Portugal/PT1806/2020\|EPI_ISL_732031\|2020-1 | ···················· | ···················· | ····················· | ···················· | ···················· | ···················· |
| Y | 5930 | hCoV-19/Portugal/PT1807/2020\|EPI_ISL_732032\|2020-1 | ···················· | ···················· | ····················· | ···················· | ···················· | ···················· |
| Y | 5931 | hCoV-19/Portugal/PT1808/2020\|EPI_ISL_732033\|2020-1 | ···················· | ···················· | ····················· | ···················· | ···················· | ···················· |
| Y | 5932 | hCoV-19/Portugal/PT1809/2020\|EPI_ISL_732034\|2020-1 | ···················· | ···················· | ····················· | ···················· | ···················· | ···················· |
| Y | 5933 | hCoV-19/Portugal/PT1810/2020\|EPI_ISL_732035\|2020-1 | ···················· | ···················· | ····················· | ···················· | ···················· | ···················· |
| Y | 5934 | hCoV-19/Portugal/PT1811/2020\|EPI_ISL_732036\|2020-1 | ···················· | ···················· | ····················· | ···················· | ···················· | ···················· |
| Y | 5935 | hCoV-19/Portugal/PT1812/2020\|EPI_ISL_732037\|2020-1 | ···················· | ···················· | ····················· | ···················· | ···················· | ···················· |
| Y | 5936 | hCoV-19/Portugal/PT1814/2020\|EPI_ISL_732038\|2020-1 | ···················· | ···················· | ····················· | ···················· | ···················· | ···················· |
| Y | 5937 | hCoV-19/Portugal/PT1816/2020\|EPI_ISL_732039\|2020-1 | ···················· | ···················· | ····················· | ···················· | ···················· | ···················· |
| Y | 5938 | hCoV-19/Portugal/PT1817/2020\|EPI_ISL_732040\|2020-1 | ···················· | ···················· | ····················· | ···················· | ···················· | ···················· |
| Y | 5939 | hCoV-19/Portugal/PT1818/2020\|EPI_ISL_732041\|2020-1 | ···················· | ···················· | ····················· | ···················· | ···················· | ···················· |
| Y | 5940 | hCoV-19/Portugal/PT1819/2020\|EPI_ISL_732042\|2020-1 | ···················· | ···················· | ····················· | ···················· | ···················· | ···················· |
| Y | 5941 | hCoV-19/Portugal/PT1821/2020\|EPI_ISL_732043\|2020-1 | ···················· | ···················· | ····················· | ···················· | ···················· | ···················· |
| Y | 5942 | hCoV-19/Portugal/PT1822/2020\|EPI_ISL_732044\|2020-1 | ···················· | ···················· | ····················· | ···················· | ···················· | ···················· |
| Y | 5943 | hCoV-19/Portugal/PT1823/2020\|EPI_ISL_732045\|2020-1 | ···················· | ···················· | ····················· | ···················· | ···················· | ···················· |
| Y | 5944 | hCoV-19/Portugal/PT1824/2020\|EPI_ISL_732046\|2020-1 | ···················· | ···················· | ····················· | ···················· | ···················· | ···················· |
| Y | 5945 | hCoV-19/Portugal/PT1825/2020\|EPI_ISL_732047\|2020-1 | ···················· | ···················· | ····················· | ···················· | ···················· | ···················· |
| Y | 5946 | hCoV-19/Portugal/PT1826/2020\|EPI_ISL_732048\|2020-1 | ···················· | ···················· | ····················· | ···················· | ···················· | ···················· |
| Y | 5947 | hCoV-19/Portugal/PT1827/2020\|EPI_ISL_732049\|2020-1 | ···················· | ···················· | ····················· | ···················· | ···················· | ···················· |
| Y | 5948 | hCoV-19/Russia/KL-RII-MH5356S/2020\|EPI_ISL_733300\| | ···················· | ···················· | ····················· | ···················· | ···················· | ···················· |
| Y | 5949 | hCoV-19/Russia/PSK-RII-MH5604S/2020\|EPI_ISL_733303 | ···················· | ···················· | ····················· | ···················· | ···················· | ···················· |
| Y | 5950 | hCoV-19/Russia/LEN-RII-MH5616S/2020\|EPI_ISL_733304 | ···················· | ···················· | ····················· | ···················· | ···················· | ···················· |
| Y | 5951 | hCoV-19/Russia/PSK-RII-MH5619S/2020\|EPI_ISL_733305 | ···················· | ···················· | ····················· | ···················· | ···················· | ···················· |
| Y | 5952 | hCoV-19/Russia/PSK-RII-MH5633S/2020\|EPI_ISL_733308 | ···················· | ···················· | ····················· | ···················· | ···················· | ···················· |
| Y | 5953 | hCoV-19/Russia/LEN-RII-MH5654S/2020\|EPI_ISL_733309 | ···················· | ···················· | ····················· | ···················· | ···················· | ···················· |
| Y | 5954 | hCoV-19/Russia/PSK-RII-MH5658S/2020\|EPI_ISL_733310 | ···················· | ···················· | ····················· | ···················· | ···················· | ···················· |
| Y | 5955 | hCoV-19/Russia/SPE-RII-MH5887S/2020\|EPI_ISL_733318 | ···················· | ···················· | ····················· | ···················· | ···················· | ···················· |
| Y | 5956 | hCoV-19/Russia/SPE-RII-MH5892S/2020\|EPI_ISL_733321 | ···················· | ···················· | ····················· | ···················· | ···················· | ···················· |
| Y | 5957 | hCoV-19/Russia/SPE-RII-MH5894S/2020\|EPI_ISL_733323 | ···················· | ···················· | ····················· | ···················· | ···················· | ···················· |
| Y | 5958 | hCoV-19/Russia/SPE-RII-MH5895S/2020\|EPI_ISL_733324 | ···················· | ···················· | ····················· | ···················· | ···················· | ···················· |
| Y | 5959 | hCoV-19/Russia/SPE-RII-MH5897S/2020\|EPI_ISL_733325 | ···················· | ···················· | ····················· | ···················· | ···················· | ···················· |
| Y | 5960 | hCoV-19/Russia/LEN-RII-MH5948S/2020\|EPI_ISL_733331 | ···················· | ···················· | ····················· | ···················· | ···················· | ···················· |
| Y | 5961 | hCoV-19/Russia/PSK-RII-MH5952S/2020\|EPI_ISL_733332 | ···················· | ···················· | ····················· | ···················· | ···················· | ···················· |
| Y | 5962 | hCoV-19/Russia/NGR-RII-MH5963S/2020\|EPI_ISL_733333 | ···················· | ···················· | ····················· | ···················· | ···················· | ···················· |
| Y | 5963 | hCoV-19/Russia/MUR-RII-MH6034S/2020\|EPI_ISL_733334 | ···················· | ···················· | ····················· | ···················· | ···················· | ···················· |
| Y | 5964 | hCoV-19/Russia/LEN-RII-MH6062S/2020\|EPI_ISL_733337 | ···················· | ···················· | ·········T··········· | ···················· | ···················· | ···················· |
| Y | 5965 | hCoV-19/Russia/PSK-RII-MH6086S/2020\|EPI_ISL_733344 | ···················· | ···················· | ····················· | ···················· | ···················· | ···················· |
| Y | 5966 | hCoV-19/Russia/PSK-RII-MH6094S/2020\|EPI_ISL_733345 | ···················· | ···················· | ····················· | ···················· | ···················· | ···················· |
| Y | 5967 | hCoV-19/Russia/NGR-RII-MH6127S/2020\|EPI_ISL_733350 | ···················· | ···················· | ····················· | ···················· | ···················· | ···················· |
| Y | 5968 | hCoV-19/Russia/LEN-RII-MH6145S/2020\|EPI_ISL_733351 | ···················· | ···················· | ····················· | ···················· | ···················· | ···················· |
| Y | 5969 | hCoV-19/Russia/PSK-RII-MH6156S/2020\|EPI_ISL_733353 | ···················· | ···················· | ····················· | ···················· | ···················· | ···················· |
| Y | 5970 | hCoV-19/Russia/PSK-RII-MH6161S/2020\|EPI_ISL_733354 | ···················· | ···················· | ····················· | ···················· | ···················· | ···················· |
| Y | 5971 | hCoV-19/Russia/MUR-RII-MH6168S/2020\|EPI_ISL_733355 | ···················· | ···················· | ····················· | ···················· | ···················· | ···················· |
| Y | 5972 | hCoV-19/Russia/PSK-RII-MH6191S/2020\|EPI_ISL_733361 | ···················· | ···················· | ····················· | ···················· | ···················· | ···················· |
| Y | 5973 | hCoV-19/Russia/MUR-RII-MH6271S/2020\|EPI_ISL_733366 | ···················· | ···················· | ····················· | ···················· | ···················· | ···················· |
| Y | 5974 | hCoV-19/Russia/PSK-RII-MH6287S/2020\|EPI_ISL_733368 | ···················· | ···················· | ····················· | ···················· | ···················· | ···················· |
| Y | 5975 | hCoV-19/Russia/NGR-RII-MH6288S/2020\|EPI_ISL_733369 | ···················· | ···················· | ····················· | ···················· | ···················· | ···················· |
| Y | 5976 | hCoV-19/Russia/PSK-RII-MH6291S/2020\|EPI_ISL_733371 | ···················· | ···················· | ····················· | ···················· | ···················· | ···················· |
| Y | 5977 | hCoV-19/Russia/NGR-RII-MH6296S/2020\|EPI_ISL_733374 | ···················· | ···················· | ····················· | ···················· | ···················· | ···················· |

| | | | | | | | | |
|---|---|---|---|---|---|---|---|---|
| Y | 5978 | hCoV-19/Russia/NGR-RII-MH6306S/2020\|EPI_ISL_733376 | .................... | .................... | .................... | .................... | .................... | .................... |
| Y | 5979 | hCoV-19/Russia/PSK-RII-MH6314S/2020\|EPI_ISL_733378 | .................... | .................... | .................... | .................... | .................... | .................... |
| Y | 5980 | hCoV-19/Russia/PSK-RII-MH6316S/2020\|EPI_ISL_733379 | .................... | .................... | .................... | .................... | .................... | .................... |
| Y | 5981 | hCoV-19/Belgium/rega-0417508/2020\|EPI_ISL_734600\|2 | .................... | .................... | .................... | .................... | .................... | .................... |
| Y | 5982 | hCoV-19/Belgium/rega-0417509/2020\|EPI_ISL_734601\|2 | .................... | .................... | .................... | .................... | .................... | .................... |
| Y | 5983 | hCoV-19/Belgium/rega-0417510/2020\|EPI_ISL_734602\|2 | .................... | .................... | .................... | .................... | .................... | .................... |
| Y | 5984 | hCoV-19/Belgium/rega-0417511/2020\|EPI_ISL_734603\|2 | .................... | .................... | .................... | .................... | .................... | .................... |
| Y | 5985 | hCoV-19/Belgium/rega-0417512/2020\|EPI_ISL_734604\|2 | .................... | .................... | .................... | .................... | .................... | .................... |
| Y | 5986 | hCoV-19/Belgium/rega-0417513/2020\|EPI_ISL_734605\|2 | .................... | .................... | .................... | .................... | .................... | .................... |
| Y | 5987 | hCoV-19/Belgium/rega-0417514/2020\|EPI_ISL_734606\|2 | .................... | .................... | .................... | .................... | .................... | .................... |
| Y | 5988 | hCoV-19/Belgium/rega-0417515/2020\|EPI_ISL_734607\|2 | .................... | .................... | .................... | .................... | .................... | .................... |
| Y | 5989 | hCoV-19/Belgium/rega-0417516/2020\|EPI_ISL_734608\|2 | .................... | .................... | .................... | .................... | .................... | .................... |
| Y | 5990 | hCoV-19/Belgium/rega-0417517/2020\|EPI_ISL_734609\|2 | .................... | .................... | .................... | .................... | .................... | .................... |
| Y | 5991 | hCoV-19/Belgium/rega-0417518/2020\|EPI_ISL_734610\|2 | .................... | .................... | .................... | .................... | .................... | .................... |
| Y | 5992 | hCoV-19/Belgium/rega-0417519/2020\|EPI_ISL_734611\|2 | .................... | .................... | .................... | .................... | .................... | .................... |
| Y | 5993 | hCoV-19/Belgium/rega-0417520/2020\|EPI_ISL_734612\|2 | .................... | .................... | .................... | .................... | .................... | .................... |
| Y | 5994 | hCoV-19/Belgium/rega-0417521/2020\|EPI_ISL_734613\|2 | .................... | .................... | .................... | .................... | .................... | .................... |
| Y | 5995 | hCoV-19/Belgium/rega-0417523/2020\|EPI_ISL_734614\|2 | .................... | .................... | .................... | .................... | .................... | .................... |
| Y | 5996 | hCoV-19/Belgium/rega-0417524/2020\|EPI_ISL_734615\|2 | .................... | .................... | .................... | .................... | .................... | .................... |
| Y | 5997 | hCoV-19/Belgium/rega-0417525/2020\|EPI_ISL_734616\|2 | .................... | .................... | .................... | .................... | .................... | .................... |
| Y | 5998 | hCoV-19/Belgium/rega-0417526/2020\|EPI_ISL_734617\|2 | .................... | .................... | .................... | .................... | .................... | .................... |
| Y | 5999 | hCoV-19/Belgium/rega-0417687/2020\|EPI_ISL_734618\|2 | .................... | .................... | .................... | .................... | .................... | .................... |
| Y | 6000 | hCoV-19/Belgium/rega-0417908/2020\|EPI_ISL_734619\|2 | .................... | .................... | .................... | .................... | .................... | .................... |
| Y | 6001 | hCoV-19/Belgium/rega-0418527/2020\|EPI_ISL_734620\|2 | .................... | .................... | .................... | .................... | .................... | .................... |
| Y | 6002 | hCoV-19/Belgium/rega-0418528/2020\|EPI_ISL_734621\|2 | .................... | .................... | .................... | .................... | .................... | .................... |
| Y | 6003 | hCoV-19/Belgium/rega-0418529/2020\|EPI_ISL_734622\|2 | .................... | .................... | .................... | .................... | .................... | .................... |
| Y | 6004 | hCoV-19/Belgium/rega-0419530/2020\|EPI_ISL_734623\|2 | .................... | .................... | .................... | .................... | .................... | .................... |
| Y | 6005 | hCoV-19/Belgium/rega-0419531/2020\|EPI_ISL_734624\|2 | .................... | .................... | .................... | .................... | .................... | .................... |
| Y | 6006 | hCoV-19/Belgium/rega-0419532/2020\|EPI_ISL_734625\|2 | .................... | .................... | .................... | .................... | .................... | .................... |
| Y | 6007 | hCoV-19/Belgium/rega-0419533/2020\|EPI_ISL_734626\|2 | .................... | .................... | .................... | .................... | .................... | .................... |
| Y | 6008 | hCoV-19/Belgium/rega-0419534/2020\|EPI_ISL_734627\|2 | .................... | .................... | .................... | .................... | .................... | .................... |
| Y | 6009 | hCoV-19/Belgium/rega-0419535/2020\|EPI_ISL_734628\|2 | .................... | .................... | .................... | .................... | .................... | .................... |
| Y | 6010 | hCoV-19/Belgium/rega-0419536/2020\|EPI_ISL_734629\|2 | .................... | .................... | .................... | .................... | .................... | .................... |
| Y | 6011 | hCoV-19/Belgium/rega-0419585/2020\|EPI_ISL_734630\|2 | .................... | .................... | .................... | .................... | .................... | .................... |
| Y | 6012 | hCoV-19/Belgium/rega-0419586/2020\|EPI_ISL_734631\|2 | .................... | .................... | .................... | .................... | .................... | .................... |
| Y | 6013 | hCoV-19/Belgium/rega-0420909/2020\|EPI_ISL_734632\|2 | .................... | .................... | .................... | .................... | .................... | .................... |
| Y | 6014 | hCoV-19/Belgium/rega-0422587/2020\|EPI_ISL_734633\|2 | .................... | .................... | .................... | .................... | .................... | .................... |
| Y | 6015 | hCoV-19/Belgium/rega-0422588/2020\|EPI_ISL_734634\|2 | .................... | .................... | .................... | .................... | .................... | .................... |
| Y | 6016 | hCoV-19/Belgium/rega-0422589/2020\|EPI_ISL_734635\|2 | .................... | .................... | .................... | .................... | .................... | .................... |
| Y | 6017 | hCoV-19/Belgium/rega-0422590/2020\|EPI_ISL_734636\|2 | .................... | .................... | .................... | .................... | .................... | .................... |
| Y | 6018 | hCoV-19/Belgium/rega-0422591/2020\|EPI_ISL_734637\|2 | .................... | .................... | .................... | .................... | .................... | .................... |
| Y | 6019 | hCoV-19/Belgium/rega-0422592/2020\|EPI_ISL_734638\|2 | .................... | .................... | .................... | .................... | .................... | .................... |
| Y | 6020 | hCoV-19/Belgium/rega-0422656/2020\|EPI_ISL_734639\|2 | .................... | .................... | .................... | .................... | .................... | .................... |
| Y | 6021 | hCoV-19/Belgium/rega-0423593/2020\|EPI_ISL_734640\|2 | .................... | .................... | .................... | .................... | .................... | .................... |
| Y | 6022 | hCoV-19/Belgium/rega-0423594/2020\|EPI_ISL_734641\|2 | .................... | .................... | .................... | .................... | .................... | .................... |
| Y | 6023 | hCoV-19/Belgium/rega-0423595/2020\|EPI_ISL_734642\|2 | .................... | .................... | .................... | .................... | .................... | .................... |
| Y | 6024 | hCoV-19/Belgium/rega-0423596/2020\|EPI_ISL_734643\|2 | .................... | .................... | .................... | .................... | .................... | .................... |
| Y | 6025 | hCoV-19/Belgium/rega-0423597/2020\|EPI_ISL_734644\|2 | .................... | .................... | .................... | .................... | .................... | .................... |
| Y | 6026 | hCoV-19/Belgium/rega-0423598/2020\|EPI_ISL_734645\|2 | .................... | .................... | .................... | .................... | .................... | .................... |
| Y | 6027 | hCoV-19/Belgium/rega-0423599/2020\|EPI_ISL_734646\|2 | .................... | .................... | .................... | .................... | .................... | .................... |
| Y | 6028 | hCoV-19/Belgium/rega-0423600/2020\|EPI_ISL_734647\|2 | .................... | .................... | .................... | .................... | .................... | .................... |
| Y | 6029 | hCoV-19/Belgium/rega-0423657/2020\|EPI_ISL_734648\|2 | .................... | .................... | .................... | .................... | .................... | .................... |
| Y | 6030 | hCoV-19/Belgium/rega-0424910/2020\|EPI_ISL_734649\|2 | .................... | .................... | .................... | .................... | .................... | .................... |
| Y | 6031 | hCoV-19/Belgium/rega-04261189/2020\|EPI_ISL_734650\| | .................... | .................... | .................... | .................... | .................... | .................... |
| Y | 6032 | hCoV-19/Belgium/rega-04261190/2020\|EPI_ISL_734651\| | .................... | .................... | .................... | .................... | .................... | .................... |
| Y | 6033 | hCoV-19/Belgium/rega-04261191/2020\|EPI_ISL_734652\| | .................... | .................... | .................... | .................... | .................... | .................... |
| Y | 6034 | hCoV-19/Belgium/rega-0426603/2020\|EPI_ISL_734653\|2 | .................... | .................... | .................... | .................... | .................... | .................... |
| Y | 6035 | hCoV-19/Belgium/rega-0426604/2020\|EPI_ISL_734654\|2 | .................... | .................... | .................... | .................... | .................... | .................... |
| Y | 6036 | hCoV-19/Belgium/rega-0426605/2020\|EPI_ISL_734655\|2 | .................... | .................... | .................... | .................... | .................... | .................... |
| Y | 6037 | hCoV-19/Belgium/rega-0426606/2020\|EPI_ISL_734656\|2 | .................... | .................... | .................... | .................... | .................... | .................... |
| Y | 6038 | hCoV-19/Belgium/rega-04271192/2020\|EPI_ISL_734657\| | .................... | .................... | .................... | .................... | .................... | .................... |
| Y | 6039 | hCoV-19/Belgium/rega-0427607/2020\|EPI_ISL_734658\|2 | .................... | .................... | .................... | .................... | .................... | .................... |
| Y | 6040 | hCoV-19/Belgium/rega-0427608/2020\|EPI_ISL_734659\|2 | .................... | .................... | .................... | .................... | .................... | .................... |
| Y | 6041 | hCoV-19/Belgium/rega-0427609/2020\|EPI_ISL_734660\|2 | .................... | .................... | .................... | .................... | .................... | .................... |
| Y | 6042 | hCoV-19/Belgium/rega-0427610/2020\|EPI_ISL_734661\|2 | .................... | .................... | .................... | .................... | .................... | .................... |
| Y | 6043 | hCoV-19/Belgium/rega-0427611/2020\|EPI_ISL_734662\|2 | .................... | .................... | .................... | .................... | .................... | .................... |
| Y | 6044 | hCoV-19/Belgium/rega-0427612/2020\|EPI_ISL_734663\|2 | .................... | .................... | .................... | .................... | .................... | .................... |
| Y | 6045 | hCoV-19/Belgium/rega-0427613/2020\|EPI_ISL_734664\|2 | .................... | .................... | .................... | .................... | .................... | .................... |
| Y | 6046 | hCoV-19/Belgium/rega-0427614/2020\|EPI_ISL_734665\|2 | .................... | .................... | .................... | .................... | .................... | .................... |
| Y | 6047 | hCoV-19/Belgium/rega-0427615/2020\|EPI_ISL_734666\|2 | .................... | .................... | .................... | .................... | .................... | .................... |
| Y | 6048 | hCoV-19/Belgium/rega-0427616/2020\|EPI_ISL_734667\|2 | .................... | .................... | .................... | .................... | .................... | .................... |
| Y | 6049 | hCoV-19/Belgium/rega-0427617/2020\|EPI_ISL_734668\|2 | .................... | .................... | .................... | .................... | .................... | .................... |
| Y | 6050 | hCoV-19/Belgium/rega-0427658/2020\|EPI_ISL_734669\|2 | .................... | .................... | .................... | .................... | .................... | .................... |
| Y | 6051 | hCoV-19/Belgium/rega-0429624/2020\|EPI_ISL_734677\|2 | .................... | .................... | .................... | .................... | .................... | .................... |
| Y | 6052 | hCoV-19/Belgium/rega-0429625/2020\|EPI_ISL_734678\|2 | .................... | .................... | .................... | .................... | .................... | .................... |
| Y | 6053 | hCoV-19/Belgium/rega-0429627/2020\|EPI_ISL_734680\|2 | .................... | .................... | .................... | .................... | .................... | .................... |
| Y | 6054 | hCoV-19/Belgium/rega-0429628/2020\|EPI_ISL_734681\|2 | .................... | .................... | .................... | .................... | .................... | .................... |
| Y | 6055 | hCoV-19/Belgium/rega-0429659/2020\|EPI_ISL_734682\|2 | .................... | .................... | .................... | .................... | .................... | .................... |
| Y | 6056 | hCoV-19/Belgium/rega-0429662/2020\|EPI_ISL_734683\|2 | .................... | .................... | .................... | .................... | .................... | .................... |
| Y | 6057 | hCoV-19/Belgium/rega-0429663/2020\|EPI_ISL_734684\|2 | .................... | .................... | .................... | .................... | .................... | .................... |

Y 6058 hCoV-19/Belgium/rega-0429665/2020|EPI_ISL_734685|2 ···················· ···················· ···················· ···················· ···················· ····················
Y 6059 hCoV-19/Belgium/rega-0429911/2020|EPI_ISL_734686|2 ···················· ···················· ···················· ···················· ···················· ····················
Y 6060 hCoV-19/Belgium/rega-0430630/2020|EPI_ISL_734687|2 ···················· ···················· ···················· ···················· ···················· ····················
Y 6061 hCoV-19/Belgium/rega-0430631/2020|EPI_ISL_734688|2 ···················· ···················· ···················· ···················· ···················· ····················
Y 6062 hCoV-19/Belgium/rega-0501632/2020|EPI_ISL_734689|2 ···················· ·A·················· ···················· ···················· ···················· ····················
Y 6063 hCoV-19/Belgium/rega-0501633/2020|EPI_ISL_734690|2 ···················· ···················· ···················· ···················· ···················· ····················
Y 6064 hCoV-19/Belgium/rega-0501634/2020|EPI_ISL_734691|2 ···················· ···················· ···················· ···················· ···················· ····················
Y 6065 hCoV-19/Belgium/rega-0501635/2020|EPI_ISL_734692|2 ···················· ···················· ···················· ···················· ···················· ····················
Y 6066 hCoV-19/Belgium/rega-0501636/2020|EPI_ISL_734693|2 ···················· ···················· ···················· ···················· ···················· ····················
Y 6067 hCoV-19/Belgium/rega-0501666/2020|EPI_ISL_734694|2 ···················· ···················· ···················· ···················· ···················· ····················
Y 6068 hCoV-19/Belgium/rega-0502637/2020|EPI_ISL_734695|2 ···················· ···················· ···················· ···················· ···················· ····················
Y 6069 hCoV-19/Belgium/rega-0502638/2020|EPI_ISL_734696|2 ···················· ···················· ···················· ···················· ···················· ····················
Y 6070 hCoV-19/Belgium/rega-0502639/2020|EPI_ISL_734697|2 ···················· ···················· ···················· ···················· ···················· ····················
Y 6071 hCoV-19/Belgium/rega-0502640/2020|EPI_ISL_734698|2 ···················· ···················· ···················· ···················· ···················· ····················
Y 6072 hCoV-19/Belgium/rega-0502900/2020|EPI_ISL_734699|2 ···················· ···················· ···················· ···················· ···················· ····················
Y 6073 hCoV-19/England/ALDP-C5837D/2020|EPI_ISL_735913|20 ···················· ···················· ···················· ···················· ···················· ····················
Y 6074 hCoV-19/England/ALDP-C57FD4/2020|EPI_ISL_735914|20 ···················· ···················· ···················· ···················· ···················· ····················
Y 6075 hCoV-19/England/ALDP-C58780/2020|EPI_ISL_735916|20 ···················· ···················· ···················· ···················· ···················· ····················
Y 6076 hCoV-19/England/QEUH-C5B85D/2020|EPI_ISL_735920|20 ···················· ···················· ···················· ···················· ···················· ····················
Y 6077 hCoV-19/England/ALDP-C59172/2020|EPI_ISL_735922|20 ···················· ···················· ···················· ···················· ···················· ····················
Y 6078 hCoV-19/England/ALDP-C58BC0/2020|EPI_ISL_735923|20 ···················· ···················· ···················· ···················· ···················· ····················
Y 6079 hCoV-19/England/ALDP-C5A01A/2020|EPI_ISL_735925|20 ···················· ···················· ···················· ···················· ···················· ····················
Y 6080 hCoV-19/England/ALDP-C5852C/2020|EPI_ISL_735926|20 ···················· ···················· ···················· ···················· ···················· ····················
Y 6081 hCoV-19/England/ALDP-C58243/2020|EPI_ISL_735928|20 ···················· ···················· ···················· ···················· ···················· ····················
Y 6082 hCoV-19/England/ALDP-C5854A/2020|EPI_ISL_735930|20 ···················· ···················· ···················· ···················· ···················· ····················
Y 6083 hCoV-19/England/ALDP-C5845C/2020|EPI_ISL_735933|20 ···················· ···················· ···················· ···················· ···················· ····················
Y 6084 hCoV-19/England/ALDP-C58F6A/2020|EPI_ISL_735936|20 ···················· ···················· ···················· ···················· ···················· ····················
Y 6085 hCoV-19/England/QEUH-C5BD4F/2020|EPI_ISL_735939|20 ···················· ···················· ···················· ···················· ···················· ····················
Y 6086 hCoV-19/England/ALDP-C588AB/2020|EPI_ISL_735941|20 ···················· ···················· ···················· ···················· ···················· ····················
Y 6087 hCoV-19/England/ALDP-C57FB6/2020|EPI_ISL_735943|20 ···················· ···················· ···················· ···················· ···················· ····················
Y 6088 hCoV-19/England/ALDP-C581EC/2020|EPI_ISL_735944|20 ···················· ···················· ···················· ···················· ···················· ····················
Y 6089 hCoV-19/England/ALDP-C58401/2020|EPI_ISL_735947|20 ···················· ···················· ···················· ···················· ···················· ····················
Y 6090 hCoV-19/England/ALDP-C58207/2020|EPI_ISL_735950|20 ···················· ···················· ···················· ···················· ···················· ····················
Y 6091 hCoV-19/England/ALDP-C58F00/2020|EPI_ISL_735951|20 ···················· ···················· ···················· ···················· ···················· ····················
Y 6092 hCoV-19/England/QEUH-C5B83F/2020|EPI_ISL_735952|20 ···················· ···················· ···················· ···················· ···················· ····················
Y 6093 hCoV-19/England/ALDP-C582DA/2020|EPI_ISL_735954|20 ···················· ···················· ···················· ···················· ···················· ····················
Y 6094 hCoV-19/England/QEUH-C5BD30/2020|EPI_ISL_735956|20 ···················· ···················· ···················· ···················· ···················· ····················
Y 6095 hCoV-19/England/ALDP-C5A00B/2020|EPI_ISL_735957|20 ···················· ···················· ···················· ···················· ···················· ····················
Y 6096 hCoV-19/England/ALDP-C57C28/2020|EPI_ISL_735958|20 ···················· ···················· ···················· ···················· ···················· ····················
Y 6097 hCoV-19/England/ALDP-C58683/2020|EPI_ISL_735959|20 ···················· ···················· ···················· ···················· ······T············· ····················
Y 6098 hCoV-19/England/QEUH-C5C7B9/2020|EPI_ISL_735961|20 ···················· ···················· ···················· ···················· ···················· ····················
Y 6099 hCoV-19/England/ALDP-C581CE/2020|EPI_ISL_735962|20 ···················· ···················· ···················· ···················· ···················· ····················
Y 6100 hCoV-19/England/ALDP-C57CDD/2020|EPI_ISL_735963|20 ···················· ···················· ···················· ···················· ···················· ····················
Y 6101 hCoV-19/England/ALDP-C57F89/2020|EPI_ISL_735964|20 ···················· ···················· ···················· ···················· ···················· ····················
Y 6102 hCoV-19/England/ALDP-C580EF/2020|EPI_ISL_735970|20 ···················· ···················· ···················· ···················· ···················· ····················
Y 6103 hCoV-19/England/QEUH-C5C75F/2020|EPI_ISL_735972|20 ···················· ···················· ···················· ···················· ···················· ····················
Y 6104 hCoV-19/England/ALDP-C58F4C/2020|EPI_ISL_735974|20 ···················· ···················· ···················· ···················· ···················· ····················
Y 6105 hCoV-19/England/ALDP-C58F2E/2020|EPI_ISL_735975|20 ···················· ···················· ···················· ···················· ···················· ····················
Y 6106 hCoV-19/England/ALDP-C57CBF/2020|EPI_ISL_735976|20 ···················· ···················· ···················· ···················· ···················· ····················
Y 6107 hCoV-19/England/QEUH-C5B88A/2020|EPI_ISL_735978|20 ···················· ···················· ···················· ···················· ···················· ····················
Y 6108 hCoV-19/England/ALDP-C57DDA/2020|EPI_ISL_735979|20 ···················· ···················· ···················· ···················· ···················· ····················
Y 6109 hCoV-19/England/ALDP-C5802B/2020|EPI_ISL_735980|20 ···················· ···················· ···················· ···················· ···················· ····················
Y 6110 hCoV-19/England/ALDP-C57CFB/2020|EPI_ISL_735981|20 ···················· ···················· ···················· ···················· ···················· ····················
Y 6111 hCoV-19/England/ALDP-C583E6/2020|EPI_ISL_735984|20 ···················· ···················· ···················· ···················· ···················· ····················
Y 6112 hCoV-19/England/ALDP-C58155/2020|EPI_ISL_735985|20 ···················· ···················· ···················· ···················· ···················· ····················
Y 6113 hCoV-19/England/ALDP-C58C09/2020|EPI_ISL_735986|20 ···················· ···················· ···················· ···················· ···················· ····················
Y 6114 hCoV-19/England/QEUH-C5B7D8/2020|EPI_ISL_735987|20 ···················· ···················· ···················· ···················· ···················· ····················
Y 6115 hCoV-19/England/ALDP-C580FE/2020|EPI_ISL_735988|20 ···················· ···················· ···················· ···················· ···················· ····················
Y 6116 hCoV-19/England/ALDP-C58CFA/2020|EPI_ISL_735989|20 ···················· ···················· ···················· ···················· ···················· ····················
Y 6117 hCoV-19/England/QEUH-C5C740/2020|EPI_ISL_735993|20 ···················· ···················· ···················· ···················· ···················· ····················
Y 6118 hCoV-19/England/QEUH-C5BCD9/2020|EPI_ISL_735995|20 ···················· ···················· ···················· ···················· ···················· ····················
Y 6119 hCoV-19/England/ALDP-C585A4/2020|EPI_ISL_735997|20 ···················· ·········T·········· ···················· ···················· ···················· ····················
Y 6120 hCoV-19/England/ALDP-C57FA7/2020|EPI_ISL_735998|20 ···················· ···················· ···················· ···················· ···················· ····················
Y 6121 hCoV-19/USA/TX-DSHS-1805/2020|EPI_ISL_738158|2020- ···················· ···················· ···················· ···················· ···················· ····················
Y 6122 hCoV-19/USA/TX-DSHS-1784/2020|EPI_ISL_738160|2020- ···················· ···················· ···················· ···················· ···················· ····················
Y 6123 hCoV-19/USA/TX-DSHS-1785/2020|EPI_ISL_738161|2020- ···················· ···················· ···················· ···················· ···················· ····················
Y 6124 hCoV-19/USA/TX-DSHS-1787/2020|EPI_ISL_738162|2020- ···················· ···················· ···················· ···················· ···················· ····················
Y 6125 hCoV-19/USA/TX-DSHS-1788/2020|EPI_ISL_738163|2020- ···················· ···················· ···················· ···················· ···················· ····················
Y 6126 hCoV-19/USA/TX-DSHS-1789/2020|EPI_ISL_738164|2020- ···················· ···················· ···················· ···················· ···················· ····················
Y 6127 hCoV-19/USA/TX-DSHS-1790/2020|EPI_ISL_738165|2020- ···················· ···················· ···················· ···················· ···················· ····················
Y 6128 hCoV-19/USA/TX-DSHS-1791/2020|EPI_ISL_738166|2020- ···················· ···················· ···················· ···················· ···················· ····················
Y 6129 hCoV-19/USA/TX-DSHS-1793/2020|EPI_ISL_738167|2020- ···················· ···················· ···················· ···················· ···················· ····················
Y 6130 hCoV-19/USA/TX-DSHS-1794/2020|EPI_ISL_738168|2020- ···················· ···················· ···················· ···················· ···················· ····················
Y 6131 hCoV-19/USA/TX-DSHS-1796/2020|EPI_ISL_738169|2020- ···················· ···················· ···················· ···················· ···················· ····················
Y 6132 hCoV-19/USA/TX-DSHS-1803/2020|EPI_ISL_738174|2020- ···················· ···················· ···················· ···················· ···················· ····················
Y 6133 hCoV-19/USA/TX-DSHS-1804/2020|EPI_ISL_738175|2020- ···················· ·A·················· ···················· ···················· ···················· ····················
Y 6134 hCoV-19/USA/TX-DSHS-1806/2020|EPI_ISL_738176|2020- ···················· ···················· ···················· ···················· ···················· ····················
Y 6135 hCoV-19/USA/TX-DSHS-1807/2020|EPI_ISL_738177|2020- ···················· ·A·················· ···················· ···················· ···················· ····················
Y 6136 hCoV-19/USA/TX-DSHS-1808/2020|EPI_ISL_738178|2020- ···················· ·············A······ ···················· ···················· ···················· ····················
Y 6137 hCoV-19/USA/TX-DSHS-1824/2020|EPI_ISL_738190|2020- ···················· ···················· ···················· ···················· ···················· ····················

| | | | | | | | | | |
|---|---|---|---|---|---|---|---|---|---|
| Y | 6138 | hCoV-19/USA/TX-DSHS-1825/2020\|EPI_ISL_738191\|2020- | ···················· | ···················· | ···················· | ···················· | ···················· | ···················· | |
| Y | 6139 | hCoV-19/USA/TX-DSHS-1827/2020\|EPI_ISL_738192\|2020- | ···················· | ·A·················· | ···················· | ···················· | ···················· | ···················· | |
| Y | 6140 | hCoV-19/Italy/TUS-C198/2020\|EPI_ISL_738194\|2020-02 | ···················· | ···················· | ···················· | ···················· | ···················· | ···················· | |
| Y | 6141 | hCoV-19/Belgium/rega-03131154/2020\|EPI_ISL_738196\| | ···················· | ···················· | ···················· | ···················· | ···················· | ···················· | |
| Y | 6142 | hCoV-19/Belgium/rega-0516680/2020\|EPI_ISL_738197\|2 | ···················· | ···················· | ···················· | ···················· | ···················· | ···················· | |
| Y | 6143 | hCoV-19/Belgium/rega-03021157/2020\|EPI_ISL_738198\| | ···················· | ···················· | ···················· | ···················· | ···················· | ···················· | |
| Y | 6144 | hCoV-19/Belgium/rega-03031158/2020\|EPI_ISL_738199\| | ···················· | ···················· | ···················· | ···················· | ···················· | ···················· | |
| Y | 6145 | hCoV-19/Wales/PHWC-4949E4/2020\|EPI_ISL_726201\|2020 | ···················· | ···················· | ···················· | ···················· | ···················· | ···················· | |
| Y | 6146 | hCoV-19/Wales/PHWC-4949F3/2020\|EPI_ISL_726202\|2020 | ···················· | ···················· | ···················· | ···················· | ···················· | ···················· | |
| Y | 6147 | hCoV-19/Wales/PHWC-494A0F/2020\|EPI_ISL_726203\|2020 | ···················· | ···················· | ···················· | ···················· | ···················· | ···················· | |
| Y | 6149 | hCoV-19/Wales/PHWC-494A3C/2020\|EPI_ISL_726205\|2020 | ···················· | ···················· | ···················· | ···················· | ···················· | ···················· | |
| Y | 6151 | hCoV-19/Wales/PHWC-494A5A/2020\|EPI_ISL_726207\|2020 | ···················· | ···················· | ···················· | ···················· | ···················· | ···················· | |
| Y | 6152 | hCoV-19/Wales/PHWC-494A78/2020\|EPI_ISL_726209\|2020 | ···················· | ······A············· | ···················· | ···················· | ···················· | ···················· | |
| Y | 6155 | hCoV-19/Wales/PHWC-494AB4/2020\|EPI_ISL_726213\|2020 | ···················· | ···················· | ···················· | ···················· | ···················· | ···················· | |
| Y | 6156 | hCoV-19/Wales/PHWC-494AC3/2020\|EPI_ISL_726214\|2020 | ···················· | ···················· | ···················· | ···················· | ···················· | ···················· | |
| Y | 6157 | hCoV-19/Wales/PHWC-494AD2/2020\|EPI_ISL_726215\|2020 | ···················· | ···················· | ···················· | ···················· | ···················· | ···················· | |
| Y | 6158 | hCoV-19/Wales/PHWC-494AE1/2020\|EPI_ISL_726216\|2020 | ···················· | ···················· | ···················· | ···················· | ···················· | ···················· | |
| Y | 6159 | hCoV-19/Wales/PHWC-494AF0/2020\|EPI_ISL_726217\|2020 | ···················· | ···················· | ···················· | ···················· | ···················· | ···················· | |
| Y | 6160 | hCoV-19/Wales/PHWC-494B0C/2020\|EPI_ISL_726218\|2020 | ···················· | ···················· | ···················· | ···················· | ···················· | ···················· | |
| Y | 6162 | hCoV-19/Wales/PHWC-494B39/2020\|EPI_ISL_726220\|2020 | ···················· | ···················· | ···················· | ···················· | ···················· | ···················· | |
| Y | 6166 | hCoV-19/Wales/PHWC-494BB1/2020\|EPI_ISL_726226\|2020 | ···················· | ···················· | ···················· | ···················· | ···················· | ···················· | |
| Y | 6168 | hCoV-19/Wales/PHWC-494BDF/2020\|EPI_ISL_726228\|2020 | ···················· | ···················· | ···················· | ···················· | ···················· | ···················· | |
| Y | 6169 | hCoV-19/Wales/PHWC-494BEE/2020\|EPI_ISL_726229\|2020 | ···················· | ···················· | ···················· | ···················· | ···················· | ···················· | |
| Y | 6170 | hCoV-19/Wales/PHWC-494BFD/2020\|EPI_ISL_726230\|2020 | ···················· | ···················· | ···················· | ···················· | ···················· | ···················· | |
| Y | 6171 | hCoV-19/Wales/PHWC-494C09/2020\|EPI_ISL_726231\|2020 | ···················· | ···················· | ···················· | ···················· | ···················· | ···················· | |
| Y | 6172 | hCoV-19/Wales/PHWC-494C18/2020\|EPI_ISL_726232\|2020 | ···················· | ···················· | ···················· | ···················· | ···················· | ···················· | |
| Y | 6173 | hCoV-19/Wales/PHWC-494C27/2020\|EPI_ISL_726233\|2020 | ···················· | ···················· | ···················· | ···················· | ···················· | ···················· | |
| Y | 6175 | hCoV-19/Wales/PHWC-494C45/2020\|EPI_ISL_726235\|2020 | ···················· | ···················· | ···················· | ···················· | ···················· | ···················· | |
| Y | 6176 | hCoV-19/Wales/PHWC-494C54/2020\|EPI_ISL_726236\|2020 | ···················· | ···················· | ···················· | ···················· | ···················· | ···················· | |
| Y | 6177 | hCoV-19/Wales/PHWC-494C63/2020\|EPI_ISL_726237\|2020 | ···················· | ···················· | ···················· | ···················· | ···················· | ···················· | |
| Y | 6178 | hCoV-19/Wales/PHWC-494C72/2020\|EPI_ISL_726238\|2020 | ···················· | ···················· | ···················· | ···················· | ···················· | ···················· | |
| Y | 6179 | hCoV-19/Wales/PHWC-494C81/2020\|EPI_ISL_726239\|2020 | ···················· | ···················· | ···················· | ···················· | ···················· | ···················· | |
| Y | 6180 | hCoV-19/Wales/PHWC-494CBE/2020\|EPI_ISL_726240\|2020 | ···················· | ···················· | ···················· | ···················· | ···················· | ···················· | |
| Y | 6181 | hCoV-19/Wales/PHWC-494CCD/2020\|EPI_ISL_726241\|2020 | ···················· | ···················· | ···················· | ···················· | ···················· | ···················· | |
| Y | 6185 | hCoV-19/Wales/PHWC-494D33/2020\|EPI_ISL_726246\|2020 | ···················· | ···················· | ···················· | ···················· | ···················· | ···················· | |
| Y | 6186 | hCoV-19/Wales/PHWC-494D42/2020\|EPI_ISL_726247\|2020 | ···················· | ···················· | ···················· | ···················· | ···················· | ···················· | |
| Y | 6187 | hCoV-19/Wales/PHWC-494D51/2020\|EPI_ISL_726248\|2020 | ···················· | ···················· | ···················· | ···················· | ···················· | ···················· | |
| Y | 6190 | hCoV-19/Wales/PHWC-494DD9/2020\|EPI_ISL_726255\|2020 | ···················· | ···················· | ···················· | ···················· | NNN················· | ···················· | 1 |
| Y | 6191 | hCoV-19/Wales/PHWC-494DE8/2020\|EPI_ISL_726256\|2020 | ···················· | ···················· | ···················· | ···················· | ···················· | ···················· | |
| Y | 6192 | hCoV-19/Wales/PHWC-494E12/2020\|EPI_ISL_726258\|2020 | ···················· | ···················· | ···················· | ···················· | ···················· | ···················· | |
| Y | 6194 | hCoV-19/Wales/PHWC-494F2E/2020\|EPI_ISL_726272\|2020 | ···················· | ···················· | ···················· | ···················· | ···················· | ···················· | |
| Y | 6196 | hCoV-19/Wales/PHWC-494F4C/2020\|EPI_ISL_726274\|2020 | ···················· | ···················· | ···················· | ···················· | ···················· | ···················· | |
| Y | 6197 | hCoV-19/Wales/PHWC-494F5B/2020\|EPI_ISL_726275\|2020 | ···················· | ···················· | ···················· | ···················· | ···················· | ···················· | |
| Y | 6202 | hCoV-19/Wales/PHWC-494FE2/2020\|EPI_ISL_726284\|2020 | ···················· | ···················· | ···················· | ···················· | ···················· | ···················· | |
| Y | 6204 | hCoV-19/Wales/PHWC-49500C/2020\|EPI_ISL_726286\|2020 | ···················· | ···················· | ···················· | ···················· | ···················· | ···················· | |
| Y | 6205 | hCoV-19/Wales/PHWC-49501B/2020\|EPI_ISL_726287\|2020 | ···················· | ···················· | ···················· | ···················· | ···················· | ···················· | |
| Y | 6209 | hCoV-19/Wales/PHWC-49A15F/2020\|EPI_ISL_727501\|2020 | ···················· | ···················· | ···················· | ···················· | ···················· | ···················· | |
| Y | 6211 | hCoV-19/Wales/PHWC-49A18C/2020\|EPI_ISL_727503\|2020 | ···················· | ···················· | ···················· | ···················· | ···················· | ···················· | |
| Y | 6212 | hCoV-19/Wales/PHWC-49A1AA/2020\|EPI_ISL_727505\|2020 | ···················· | ···················· | ···················· | ···················· | ···················· | ···················· | |
| Y | 6213 | hCoV-19/Wales/PHWC-49A1B9/2020\|EPI_ISL_727506\|2020 | ···················· | ···················· | ···················· | ···················· | ···················· | ···················· | |
| Y | 6214 | hCoV-19/Wales/PHWC-49A1C8/2020\|EPI_ISL_727507\|2020 | ···················· | ···················· | ···················· | ···················· | ···················· | ···················· | |
| Y | 6215 | hCoV-19/Wales/PHWC-49A1D7/2020\|EPI_ISL_727508\|2020 | ···················· | ···················· | ···················· | ···················· | ···················· | ···················· | |
| Y | 6216 | hCoV-19/Wales/PHWC-49A1E6/2020\|EPI_ISL_727509\|2020 | ···················· | ···················· | ···················· | ···················· | ···················· | ···················· | |
| Y | 6217 | hCoV-19/Wales/PHWC-49A1F5/2020\|EPI_ISL_727510\|2020 | ···················· | ···················· | ···················· | ···················· | ···················· | ···················· | |
| Y | 6218 | hCoV-19/Wales/PHWC-49A201/2020\|EPI_ISL_727511\|2020 | ···················· | ···················· | ···················· | ···················· | ···················· | ···················· | |
| Y | 6226 | hCoV-19/Wales/PHWC-49A289/2020\|EPI_ISL_727519\|2020 | ···················· | ···················· | ···················· | ···················· | ···················· | ···················· | |
| Y | 6227 | hCoV-19/Wales/PHWC-49A298/2020\|EPI_ISL_727520\|2020 | ···················· | ···················· | ···················· | ···················· | ···················· | ···················· | |
| Y | 6231 | hCoV-19/Wales/PHWC-49A2E3/2020\|EPI_ISL_727525\|2020 | ···················· | ···················· | ···················· | ···················· | ···················· | ···················· | |
| Y | 6232 | hCoV-19/Wales/PHWC-49A2F2/2020\|EPI_ISL_727526\|2020 | ···················· | ······A············· | ···················· | ···················· | ···················· | ···················· | |
| Y | 6233 | hCoV-19/Wales/PHWC-49A30E/2020\|EPI_ISL_727527\|2020 | ···················· | ···················· | ···················· | ···················· | ···················· | ···················· | |
| Y | 6235 | hCoV-19/Wales/PHWC-49A32C/2020\|EPI_ISL_727529\|2020 | ···················· | ···················· | ···················· | ···················· | ···················· | ···················· | |
| Y | 6236 | hCoV-19/Wales/PHWC-49A33B/2020\|EPI_ISL_727530\|2020 | ···················· | ···················· | ···················· | ···················· | ···················· | ···················· | |
| Y | 6240 | hCoV-19/Wales/PHWC-49A377/2020\|EPI_ISL_727534\|2020 | ···················· | ···················· | ···················· | ···················· | ···················· | ···················· | |
| Y | 6241 | hCoV-19/Wales/PHWC-49A386/2020\|EPI_ISL_727535\|2020 | ···················· | ···················· | ···················· | ···················· | ···················· | ···················· | |
| Y | 6243 | hCoV-19/Wales/PHWC-49A3B3/2020\|EPI_ISL_727537\|2020 | ···················· | ···················· | ···················· | ···················· | ···················· | ···················· | |
| Y | 6244 | hCoV-19/Wales/PHWC-49A3C2/2020\|EPI_ISL_727538\|2020 | ···················· | ···················· | ···················· | ···················· | ···················· | ···················· | |
| Y | 6245 | hCoV-19/Wales/PHWC-49A3D1/2020\|EPI_ISL_727539\|2020 | ···················· | ···················· | ···················· | ···················· | ···················· | ···················· | |
| Y | 6246 | hCoV-19/Wales/PHWC-49A3E0/2020\|EPI_ISL_727540\|2020 | ···················· | ···················· | ···················· | ···················· | ···················· | ···················· | |
| Y | 6247 | hCoV-19/Wales/PHWC-49A3FF/2020\|EPI_ISL_727541\|2020 | ···················· | ···················· | ···················· | ···················· | ···················· | ···················· | |
| Y | 6248 | hCoV-19/Wales/PHWC-49A40B/2020\|EPI_ISL_727542\|2020 | ···················· | ···················· | ···················· | ···················· | ···················· | ···················· | |
| Y | 6249 | hCoV-19/Wales/PHWC-49A41A/2020\|EPI_ISL_727543\|2020 | ···················· | ···················· | ···················· | ···················· | ···················· | ···················· | |
| Y | 6250 | hCoV-19/Wales/PHWC-49A429/2020\|EPI_ISL_727544\|2020 | ···················· | ···················· | ···················· | ···················· | ···················· | ···················· | |
| Y | 6252 | hCoV-19/Wales/PHWC-49A447/2020\|EPI_ISL_727546\|2020 | ···················· | ···················· | ···················· | ···················· | ···················· | ···················· | |
| Y | 6253 | hCoV-19/Wales/PHWC-49A465/2020\|EPI_ISL_727548\|2020 | ···················· | ······R············· | ···················· | ···················· | ···················· | ···················· | |
| Y | 6255 | hCoV-19/Wales/PHWC-49A483/2020\|EPI_ISL_727550\|2020 | ···················· | ···················· | ···················· | ···················· | ···················· | ···················· | |
| Y | 6256 | hCoV-19/Wales/PHWC-49A4B0/2020\|EPI_ISL_727553\|2020 | ···················· | ···················· | ···················· | ···················· | ···················· | ···················· | |
| Y | 6257 | hCoV-19/Wales/PHWC-49A4CF/2020\|EPI_ISL_727554\|2020 | ···················· | ···················· | ···················· | ···················· | ···················· | ···················· | |
| Y | 6258 | hCoV-19/Wales/PHWC-49A4DE/2020\|EPI_ISL_727555\|2020 | ···················· | ···················· | ···················· | ···················· | ···················· | ···················· | |
| Y | 6259 | hCoV-19/Wales/PHWC-49A4ED/2020\|EPI_ISL_727556\|2020 | ···················· | ···················· | ···················· | ···················· | ···················· | ···················· | |
| Y | 6260 | hCoV-19/Wales/PHWC-49A4FC/2020\|EPI_ISL_727557\|2020 | ···················· | ···················· | ···················· | ···················· | ···················· | ···················· | |

| | | | | | | | |
|---|---|---|---|---|---|---|---|
| Y | 6261 hCoV-19/Wales/PHWC-49A517/2020\|EPI_ISL_727558\|2020 | ···················· | ···················· | ···················· | ···················· | ···················· | ···················· |
| Y | 6262 hCoV-19/Wales/PHWC-49A526/2020\|EPI_ISL_727559\|2020 | ···················· | ···················· | ···················· | ···················· | ···················· | ···················· |
| Y | 6263 hCoV-19/Wales/PHWC-49A535/2020\|EPI_ISL_727560\|2020 | ···················· | ···················· | ···················· | ···················· | ···················· | ···················· |
| Y | 6264 hCoV-19/Wales/PHWC-49A544/2020\|EPI_ISL_727561\|2020 | ···················· | ···················· | ···················· | ···················· | ···················· | ···················· |
| Y | 6265 hCoV-19/Wales/PHWC-49A562/2020\|EPI_ISL_727563\|2020 | ···················· | ···················· | ···················· | ···················· | ···················· | ···················· |
| Y | 6266 hCoV-19/Wales/PHWC-49A571/2020\|EPI_ISL_727564\|2020 | ···················· | ···················· | ···················· | ···················· | ···················· | ···················· |
| Y | 6267 hCoV-19/Wales/PHWC-49A580/2020\|EPI_ISL_727565\|2020 | ···················· | ···················· | ···················· | ···················· | ···················· | ···················· |
| Y | 6268 hCoV-19/Wales/PHWC-49A59F/2020\|EPI_ISL_727566\|2020 | ···················· | ···················· | ···················· | ···················· | ···················· | ···················· |
| Y | 6269 hCoV-19/Wales/PHWC-49A5AE/2020\|EPI_ISL_727567\|2020 | ···················· | ···················· | ···················· | ···················· | ···················· | ···················· |
| Y | 6270 hCoV-19/Wales/PHWC-49A5BD/2020\|EPI_ISL_727568\|2020 | ···················· | ···················· | ···················· | ···················· | ···················· | ···················· |
| Y | 6271 hCoV-19/Wales/PHWC-49A5CC/2020\|EPI_ISL_727569\|2020 | ···················· | ···················· | ···················· | ···················· | ···················· | ···················· |
| Y | 6272 hCoV-19/Wales/PHWC-49A5DB/2020\|EPI_ISL_727570\|2020 | ···················· | ···················· | ···················· | ···················· | ···················· | ···················· |
| Y | 6273 hCoV-19/Wales/PHWC-49A5EA/2020\|EPI_ISL_727571\|2020 | ···················· | ···················· | ···················· | ···················· | ···················· | ···················· |
| Y | 6274 hCoV-19/Wales/PHWC-49A5F9/2020\|EPI_ISL_727572\|2020 | ···················· | ···················· | ···················· | ···················· | ···················· | ···················· |
| Y | 6275 hCoV-19/Wales/PHWC-49A605/2020\|EPI_ISL_727573\|2020 | ···················· | ···················· | ···················· | ···················· | ···················· | ···················· |
| Y | 6276 hCoV-19/Wales/PHWC-49A623/2020\|EPI_ISL_727574\|2020 | ···················· | ···················· | ···················· | ···················· | ···················· | ···················· |
| Y | 6277 hCoV-19/Wales/PHWC-49A632/2020\|EPI_ISL_727575\|2020 | ···················· | ···················· | ···················· | ···················· | ···················· | ···················· |
| Y | 6278 hCoV-19/Wales/PHWC-49A641/2020\|EPI_ISL_727576\|2020 | ···················· | ···················· | ···················· | ···················· | ···················· | ···················· |
| Y | 6279 hCoV-19/Wales/PHWC-49A650/2020\|EPI_ISL_727577\|2020 | ···················· | ···················· | ···················· | ···················· | ···················· | ···················· |
| Y | 6280 hCoV-19/Wales/PHWC-49A66F/2020\|EPI_ISL_727578\|2020 | ···················· | ···················· | ···················· | ···················· | ···················· | ···················· |
| Y | 6281 hCoV-19/Wales/PHWC-49A67E/2020\|EPI_ISL_727579\|2020 | ···················· | ···················· | ···················· | ···················· | ···················· | ···················· |
| Y | 6282 hCoV-19/Wales/PHWC-49A6AB/2020\|EPI_ISL_727581\|2020 | ···················· | ···················· | ···················· | ···················· | ···················· | ···················· |
| Y | 6284 hCoV-19/Wales/PHWC-49A6C9/2020\|EPI_ISL_727583\|2020 | ···················· | ···················· | ···················· | ···················· | ···················· | ···················· |
| Y | 6285 hCoV-19/Wales/PHWC-49A6D8/2020\|EPI_ISL_727584\|2020 | ···················· | ···················· | ···················· | ···················· | ···················· | ···················· |
| Y | 6286 hCoV-19/Wales/PHWC-49A6E7/2020\|EPI_ISL_727585\|2020 | ···················· | ···················· | ···················· | ···················· | ···················· | ···················· |
| Y | 6287 hCoV-19/Wales/PHWC-49A6F6/2020\|EPI_ISL_727586\|2020 | ···················· | ······A············· | ···················· | ···················· | ···················· | ···················· |
| Y | 6288 hCoV-19/Wales/PHWC-49A711/2020\|EPI_ISL_727587\|2020 | ···················· | ···················· | ···················· | ···················· | ···················· | ···················· |
| Y | 6289 hCoV-19/Wales/PHWC-49A720/2020\|EPI_ISL_727588\|2020 | ···················· | ···················· | ···················· | ···················· | ···················· | ···················· |
| Y | 6291 hCoV-19/Switzerland/BE-ETHZ-400339/2020\|EPI_ISL_72 | ···················· | ···················· | ···················· | ···················· | ···················· | ···················· |
| Y | 6292 hCoV-19/Switzerland/AG-ETHZ-400388/2020\|EPI_ISL_72 | ···················· | ···················· | ···················· | ···················· | ···················· | ···················· |
| Y | 6293 hCoV-19/Portugal/PT1781/2020\|EPI_ISL_732010\|2020-1 | ···················· | ···················· | ···················· | ···················· | ···················· | ···················· |
| Y | 6294 hCoV-19/Portugal/PT1782/2020\|EPI_ISL_732011\|2020-1 | ···················· | ···················· | ···················· | ···················· | ···················· | ···················· |
| Y | 6295 hCoV-19/Portugal/PT1783/2020\|EPI_ISL_732012\|2020-1 | ···················· | ···················· | ···················· | ···················· | ···················· | ···················· |
| Y | 6296 hCoV-19/Portugal/PT1784/2020\|EPI_ISL_732013\|2020-1 | ···················· | ···················· | ···················· | ···················· | ···················· | ···················· |
| Y | 6297 hCoV-19/Portugal/PT1786/2020\|EPI_ISL_732015\|2020-1 | ···················· | ···················· | ···················· | ···················· | ···················· | ···················· |
| Y | 6298 hCoV-19/Portugal/PT1787/2020\|EPI_ISL_732016\|2020-1 | ···················· | ···················· | ···················· | ···················· | ···················· | ···················· |
| Y | 6299 hCoV-19/Portugal/PT1788/2020\|EPI_ISL_732017\|2020-1 | ···················· | ···················· | ···················· | ···················· | ···················· | ···················· |
| Y | 6300 hCoV-19/Portugal/PT1789/2020\|EPI_ISL_732018\|2020-1 | ···················· | ···················· | ···················· | ···················· | ···················· | ···················· |
| Y | 6301 hCoV-19/Portugal/PT1828/2020\|EPI_ISL_732050\|2020-1 | ···················· | ···················· | ···················· | ···················· | ···················· | ···················· |
| Y | 6302 hCoV-19/Portugal/PT1829/2020\|EPI_ISL_732051\|2020-1 | ···················· | ···················· | ···················· | ···················· | ···················· | ···················· |
| Y | 6303 hCoV-19/Portugal/PT1831/2020\|EPI_ISL_732052\|2020-1 | ···················· | ···················· | ···················· | ···················· | ···················· | ···················· |
| Y | 6304 hCoV-19/Portugal/PT1832/2020\|EPI_ISL_732053\|2020-1 | ···················· | ···················· | ···················· | ···················· | ···················· | ···················· |
| Y | 6305 hCoV-19/Portugal/PT1833/2020\|EPI_ISL_732054\|2020-1 | ···················· | ···················· | ···················· | ···················· | ···················· | ···················· |
| Y | 6306 hCoV-19/Portugal/PT1834/2020\|EPI_ISL_732055\|2020-1 | ···················· | ···················· | ···················· | ···················· | ···················· | ···················· |
| Y | 6307 hCoV-19/Portugal/PT1835/2020\|EPI_ISL_732056\|2020-1 | ···················· | ···················· | ···················· | ···················· | ···················· | ···················· |
| Y | 6308 hCoV-19/Portugal/PT1837/2020\|EPI_ISL_732057\|2020-1 | ···················· | ···················· | ···················· | ···················· | ···················· | ···················· |
| Y | 6309 hCoV-19/Portugal/PT1838/2020\|EPI_ISL_732058\|2020-1 | ···················· | ···················· | ···················· | ···················· | ···················· | ···················· |
| Y | 6310 hCoV-19/Portugal/PT1839/2020\|EPI_ISL_732059\|2020-1 | ···················· | ···················· | ···················· | ···················· | ···················· | ···················· |
| Y | 6311 hCoV-19/Portugal/PT1840/2020\|EPI_ISL_732060\|2020-1 | ···················· | ···················· | ···················· | ···················· | ···················· | ···················· |
| Y | 6312 hCoV-19/Portugal/PT1841/2020\|EPI_ISL_732061\|2020-1 | ···················· | ·········A·········· | ···················· | ···················· | ······T············· | ···················· |
| Y | 6313 hCoV-19/Portugal/PT1842/2020\|EPI_ISL_732062\|2020-1 | ···················· | ···················· | ···················· | ···················· | ···················· | ···················· |
| Y | 6314 hCoV-19/Portugal/PT1843/2020\|EPI_ISL_732063\|2020-1 | ···················· | ···················· | ···················· | ···················· | ···················· | ···················· |
| Y | 6315 hCoV-19/Portugal/PT1844/2020\|EPI_ISL_732064\|2020-1 | ···················· | ···················· | ···················· | ···················· | ···················· | ···················· |
| Y | 6316 hCoV-19/Portugal/PT1845/2020\|EPI_ISL_732065\|2020-1 | ···················· | ···················· | ···················· | ···················· | ···················· | ···················· |
| Y | 6317 hCoV-19/Portugal/PT1846/2020\|EPI_ISL_732066\|2020-1 | ···················· | ···················· | ···················· | ···················· | ···················· | ···················· |
| Y | 6318 hCoV-19/Portugal/PT1847/2020\|EPI_ISL_732067\|2020-1 | ···················· | ···················· | ···················· | ···················· | ···················· | ···················· |
| Y | 6319 hCoV-19/Portugal/PT1848/2020\|EPI_ISL_732068\|2020-1 | ···················· | ···················· | ···················· | ···················· | ···················· | ···················· |
| Y | 6320 hCoV-19/Portugal/PT1849/2020\|EPI_ISL_732069\|2020-1 | ···················· | ···················· | ···················· | ···················· | ···················· | ···················· |
| Y | 6321 hCoV-19/Portugal/PT1851/2020\|EPI_ISL_732070\|2020-1 | ···················· | ···················· | ···················· | ···················· | ···················· | ···················· |
| Y | 6322 hCoV-19/Portugal/PT1852/2020\|EPI_ISL_732071\|2020-1 | ···················· | ···················· | ···················· | ···················· | ···················· | ···················· |
| Y | 6323 hCoV-19/Portugal/PT1853/2020\|EPI_ISL_732072\|2020-1 | ···················· | ···················· | ···················· | ···················· | ···················· | ···················· |
| Y | 6324 hCoV-19/Portugal/PT1854/2020\|EPI_ISL_732073\|2020-1 | ···················· | ···················· | ···················· | ···················· | ···················· | ···················· |
| Y | 6325 hCoV-19/Portugal/PT1855/2020\|EPI_ISL_732074\|2020-1 | ···················· | ···················· | ···················· | ···················· | ···················· | ···················· |
| Y | 6326 hCoV-19/Portugal/PT1856/2020\|EPI_ISL_732075\|2020-1 | ···················· | ···················· | ···················· | ···················· | ···················· | ···················· |
| Y | 6327 hCoV-19/Portugal/PT1857/2020\|EPI_ISL_732076\|2020-1 | ···················· | ···················· | ···················· | ···················· | ···················· | ···················· |
| Y | 6328 hCoV-19/Portugal/PT1858/2020\|EPI_ISL_732077\|2020-1 | ···················· | ···················· | ···················· | ···················· | ···················· | ···················· |
| Y | 6329 hCoV-19/Portugal/PT1859/2020\|EPI_ISL_732078\|2020-1 | ···················· | ···················· | ···················· | ···················· | ···················· | ···················· |
| Y | 6330 hCoV-19/Portugal/PT1860/2020\|EPI_ISL_732079\|2020-1 | ···················· | ···················· | ···················· | ···················· | ···················· | ···················· |
| Y | 6331 hCoV-19/Portugal/PT1861/2020\|EPI_ISL_732080\|2020-1 | ···················· | ···················· | ···················· | ···················· | ···················· | ···················· |
| Y | 6332 hCoV-19/Portugal/PT1862/2020\|EPI_ISL_732081\|2020-1 | ···················· | ···················· | ···················· | ···················· | ···················· | ···················· |
| Y | 6333 hCoV-19/Portugal/PT1863/2020\|EPI_ISL_732082\|2020-1 | ···················· | ···················· | ···················· | ···················· | ···················· | ···················· |
| Y | 6334 hCoV-19/Portugal/PT1864/2020\|EPI_ISL_732083\|2020-1 | ···················· | ···················· | ···················· | ···················· | ···················· | ···················· |
| Y | 6335 hCoV-19/Portugal/PT1865/2020\|EPI_ISL_732084\|2020-1 | ···················· | ···················· | ···················· | ···················· | ···················· | ···················· |
| Y | 6336 hCoV-19/Portugal/PT1866/2020\|EPI_ISL_732085\|2020-1 | ···················· | ···················· | ···················· | ···················· | ···················· | ···················· |
| Y | 6337 hCoV-19/Portugal/PT1867/2020\|EPI_ISL_732086\|2020-1 | ···················· | ···················· | ···················· | ···················· | ···················· | ···················· |
| Y | 6338 hCoV-19/Portugal/PT1868/2020\|EPI_ISL_732087\|2020-1 | ···················· | ···················· | ···················· | ···················· | ···················· | ···················· |
| Y | 6339 hCoV-19/Portugal/PT1869/2020\|EPI_ISL_732088\|2020-1 | ···················· | ···················· | ···················· | ···················· | ···················· | ···················· |
| Y | 6340 hCoV-19/Portugal/PT1870/2020\|EPI_ISL_732089\|2020-1 | ···················· | ···················· | ···················· | ···················· | ···················· | ···················· |
| Y | 6341 hCoV-19/Portugal/PT1871/2020\|EPI_ISL_732090\|2020-1 | ···················· | ···················· | ···················· | ···················· | ···················· | ···················· |
| Y | 6342 hCoV-19/Portugal/PT1872/2020\|EPI_ISL_732091\|2020-1 | ···················· | ···················· | ···················· | ···················· | ···················· | ···················· |

| | | | | | | | | |
|---|---|---|---|---|---|---|---|---|
| Y | 6343 hCoV-19/Portugal/PT1873/2020\|EPI_ISL_732092\|2020-1 | ···················· | ···················· | ···················· | ···················· | ···················· | ···················· | |
| Y | 6344 hCoV-19/Portugal/PT1875/2020\|EPI_ISL_732093\|2020-1 | ···················· | ···················· | ···················· | ···················· | ···················· | ···················· | |
| Y | 6345 hCoV-19/Portugal/PT1877/2020\|EPI_ISL_732094\|2020-1 | ···················· | ···················· | ···················· | ···················· | ···················· | ···················· | |
| Y | 6346 hCoV-19/Portugal/PT1878/2020\|EPI_ISL_732095\|2020-1 | ···················· | ···················· | ···················· | ···················· | ···················· | ···················· | |
| Y | 6347 hCoV-19/Portugal/PT1879/2020\|EPI_ISL_732096\|2020-1 | ···················· | ···················· | ···················· | ···················· | ···················· | ···················· | |
| Y | 6348 hCoV-19/Portugal/PT1880/2020\|EPI_ISL_732097\|2020-1 | ···················· | ···················· | ···················· | ···················· | ···················· | ···················· | |
| Y | 6349 hCoV-19/Portugal/PT1883/2020\|EPI_ISL_732098\|2020-1 | ···················· | ···················· | ···················· | ···················· | ···················· | ···················· | |
| Y | 6350 hCoV-19/Portugal/PT1884/2020\|EPI_ISL_732099\|2020-1 | ···················· | ···················· | ···················· | ···················· | ···················· | ···················· | |
| Y | 6351 hCoV-19/Russia/PSK-RII-MH6079S/2020\|EPI_ISL_733341 | ···················· | ···················· | ···················· | ···················· | ···················· | ···················· | |
| Y | 6352 hCoV-19/Russia/PSK-RII-MH6081S/2020\|EPI_ISL_733342 | ···················· | ···················· | ···················· | ···················· | ···················· | ···················· | |
| Y | 6353 hCoV-19/Russia/PSK-RII-MH6083S/2020\|EPI_ISL_733343 | ···················· | ···················· | ···················· | ···················· | ···················· | ···················· | |
| Y | 6354 hCoV-19/Russia/NGR-RII-MH6321S/2020\|EPI_ISL_733381 | ···················· | ···················· | ···················· | ···················· | ···················· | ···················· | |
| Y | 6355 hCoV-19/Russia/NGR-RII-MH6330S/2020\|EPI_ISL_733382 | ···················· | ···················· | ···················· | ···················· | ···················· | ···················· | |
| Y | 6356 hCoV-19/Russia/PSK-RII-MH6331S/2020\|EPI_ISL_733383 | ···················· | ···················· | ···················· | ···················· | ···················· | ···················· | |
| Y | 6357 hCoV-19/Russia/MUR-RII-MH6348S/2020\|EPI_ISL_733387 | ···················· | ···················· | ···················· | ···················· | ···················· | ···················· | |
| Y | 6358 hCoV-19/Russia/NVS-RII-MH1159V/2020\|EPI_ISL_733392 | ···················· | ···················· | ···················· | ···················· | ···················· | ···················· | |
| Y | 6359 hCoV-19/Russia/MOW-RII-MH1679V/2020\|EPI_ISL_733393 | ···················· | ···················· | ···················· | ···················· | ···················· | ···················· | |
| Y | 6360 hCoV-19/Russia/CE-RII-MH2523V/2020\|EPI_ISL_733394\| | ···················· | ···················· | ···················· | ···················· | ···················· | ···················· | |
| Y | 6361 hCoV-19/Russia/CE-RII-MH2636V/2020\|EPI_ISL_733395\| | ···················· | ···················· | ···················· | ···················· | ···················· | ···················· | |
| Y | 6362 hCoV-19/Russia/STA-RII-MH2776V/2020\|EPI_ISL_733396 | ···················· | ···················· | ···················· | ···················· | ···················· | ···················· | |
| Y | 6363 hCoV-19/Belgium/rega-04281188/2020\|EPI_ISL_734670\| | ···················· | ···················· | ···················· | ···················· | ···················· | ···················· | |
| Y | 6364 hCoV-19/Belgium/rega-0429618/2020\|EPI_ISL_734671\|2 | ···················· | ···················· | ···················· | ···················· | ···················· | ···················· | |
| Y | 6365 hCoV-19/Belgium/rega-0429619/2020\|EPI_ISL_734672\|2 | ···················· | ···················· | ···················· | ···················· | ···················· | ···················· | |
| Y | 6366 hCoV-19/Belgium/rega-0429620/2020\|EPI_ISL_734673\|2 | ···················· | ···················· | ···················· | ···················· | ···················· | ···················· | |
| Y | 6367 hCoV-19/Belgium/rega-0429621/2020\|EPI_ISL_734674\|2 | ···················· | ···················· | ···················· | ···················· | ···················· | ···················· | |
| Y | 6368 hCoV-19/Belgium/rega-0429622/2020\|EPI_ISL_734675\|2 | ···················· | ···················· | ···················· | ···················· | ···················· | ···················· | |
| Y | 6369 hCoV-19/Belgium/rega-0429623/2020\|EPI_ISL_734676\|2 | ···················· | ···················· | ···················· | ···················· | ···················· | ···················· | |
| Y | 6370 hCoV-19/Belgium/rega-0429626/2020\|EPI_ISL_734679\|2 | ···················· | ···················· | ···················· | ···················· | ···················· | ···················· | |
| Y | 6371 hCoV-19/Belgium/rega-03031160/2020\|EPI_ISL_738200\| | ···················· | ···················· | ···················· | ···················· | ···················· | ···················· | |
| Y | 6372 hCoV-19/Belgium/rega-03041161/2020\|EPI_ISL_738201\| | ···················· | ···················· | ···················· | ···················· | ···················· | ···················· | |
| Y | 6373 hCoV-19/Belgium/rega-03051162/2020\|EPI_ISL_738202\| | ···················· | ···················· | ···················· | ···················· | ···················· | ···················· | |
| Y | 6374 hCoV-19/Belgium/rega-03061153/2020\|EPI_ISL_738203\| | ···················· | ···················· | ···················· | ···················· | ···················· | ···················· | |
| Y | 6375 hCoV-19/Belgium/rega-03071156/2020\|EPI_ISL_738204\| | ···················· | ···················· | ···················· | ···················· | ···················· | ···················· | |
| Y | 6376 hCoV-19/Belgium/rega-03071159/2020\|EPI_ISL_738205\| | ···················· | ···················· | ···················· | ···················· | ···················· | ···················· | |
| Y | 6377 hCoV-19/Belgium/rega-03121152/2020\|EPI_ISL_738206\| | ···················· | ···················· | ···················· | ···················· | ···················· | ···················· | |
| Y | 6378 hCoV-19/Belgium/rega-03131155/2020\|EPI_ISL_738207\| | ···················· | ···················· | ···················· | ···················· | ···················· | ···················· | |
| Y | 6379 hCoV-19/Belgium/rega-0409682/2020\|EPI_ISL_738208\|2 | ···················· | ···················· | ···················· | ···················· | ···················· | ···················· | |
| Y | 6380 hCoV-19/Belgium/rega-0409683/2020\|EPI_ISL_738209\|2 | ···················· | ···················· | ···················· | ···················· | ···················· | ···················· | |
| Y | 6381 hCoV-19/Belgium/rega-0411672/2020\|EPI_ISL_738210\|2 | ···················· | ···················· | ···················· | ···················· | ···················· | ···················· | |
| Y | 6382 hCoV-19/Belgium/rega-0414676/2020\|EPI_ISL_738211\|2 | ···················· | ···················· | ···················· | ···················· | ···················· | ···················· | |
| Y | 6383 hCoV-19/Belgium/rega-0414677/2020\|EPI_ISL_738212\|2 | ···················· | ···················· | ···················· | ···················· | ···················· | ···················· | |
| Y | 6384 hCoV-19/Belgium/rega-0414678/2020\|EPI_ISL_738213\|2 | ···················· | ···················· | ···················· | ···················· | ···················· | ···················· | |
| Y | 6385 hCoV-19/Belgium/rega-0508673/2020\|EPI_ISL_738214\|2 | ···················· | ···················· | ···················· | ···················· | ···················· | ···················· | |
| Y | 6386 hCoV-19/Belgium/rega-0508674/2020\|EPI_ISL_738215\|2 | ···················· | ···················· | ···················· | ···················· | ···················· | ···················· | |
| Y | 6387 hCoV-19/Belgium/rega-0512675/2020\|EPI_ISL_738216\|2 | ···················· | ···················· | ···················· | ···················· | ···················· | ···················· | |
| Y | 6388 hCoV-19/Belgium/rega-0521681/2020\|EPI_ISL_738217\|2 | ···················· | ···················· | ···················· | ···················· | ···················· | ···················· | |
| Y | 6389 hCoV-19/Belgium/rega-0604718/2020\|EPI_ISL_738218\|2 | ···················· | ···················· | ···················· | ···················· | ···················· | ···················· | |
| Y | 6390 hCoV-19/Belgium/rega-0607716/2020\|EPI_ISL_738219\|2 | ···················· | ···················· | ···················· | ···················· | ···················· | ···················· | |
| Y | 6391 hCoV-19/Belgium/rega-0617717/2020\|EPI_ISL_738220\|2 | ···················· | ···················· | ···················· | ···················· | ···················· | ···················· | |
| Y | 6392 hCoV-19/Belgium/rega-0710714/2020\|EPI_ISL_738221\|2 | ···················· | ···················· | ···················· | ···················· | ···················· | ···················· | |
| Y | 6393 hCoV-19/Belgium/rega-0712715/2020\|EPI_ISL_738222\|2 | ···················· | ···················· | ···················· | ···················· | ···················· | ···················· | |
| Y | 6394 hCoV-19/Belgium/rega-09151142/2020\|EPI_ISL_738223\| | ···················· | ···················· | ···················· | ···················· | ···················· | ···················· | |
| Y | 6395 hCoV-19/Belgium/rega-09171137/2020\|EPI_ISL_738224\| | ···················· | ···················· | ···················· | ···················· | ···················· | ···················· | |
| Y | 6396 hCoV-19/Belgium/rega-09171138/2020\|EPI_ISL_738225\| | ···················· | ···················· | ···················· | ···················· | ···················· | ···················· | |
| Y | 6397 hCoV-19/Belgium/rega-09171141/2020\|EPI_ISL_738226\| | ···················· | ···················· | ···················· | ···················· | ···················· | ···················· | |
| Y | 6398 hCoV-19/Belgium/rega-09181143/2020\|EPI_ISL_738227\| | ···················· | ···················· | ···················· | ···················· | ···················· | ···················· | |
| Y | 6399 hCoV-19/Belgium/rega-09191211/2020\|EPI_ISL_738228\| | ···················· | ···················· | ···················· | ···················· | ···················· | ···················· | |
| Y | 6400 hCoV-19/Belgium/rega-09201144/2020\|EPI_ISL_738229\| | ···················· | ···················· | ···················· | ···················· | ···················· | ···················· | |
| Y | 6401 hCoV-19/Wales/PHWC-494D8E/2020\|EPI_ISL_726251\|2020 | ···················· | ···················· | ···················· | ···················· | ···················· | ···················· | |
| Y | 6405 hCoV-19/Wales/PHWC-494E6D/2020\|EPI_ISL_726263\|2020 | ···················· | ···················· | ···················· | ···················· | ···················· | ···················· | |
| Y | 6406 hCoV-19/Wales/PHWC-494EA9/2020\|EPI_ISL_726266\|2020 | ···················· | ···················· | ···················· | ···················· | ···················· | ···················· | |
| Y | 6407 hCoV-19/Wales/PHWC-494EB8/2020\|EPI_ISL_726267\|2020 | ···················· | ···················· | ···················· | ···················· | ···················· | NN·················· | |
| Y | 6413 hCoV-19/Wales/PHWC-495093/2020\|EPI_ISL_726295\|2020 | ···················· | ···················· | ···················· | ···················· | ···················· | ···················· | |
| Y | 6415 hCoV-19/Wales/PHWC-4950B1/2020\|EPI_ISL_726297\|2020 | ···················· | ···················· | ···················· | ···················· | ···················· | ···················· | |
| Y | 6416 hCoV-19/Wales/PHWC-4950C0/2020\|EPI_ISL_726298\|2020 | ···················· | ···················· | ···················· | ···················· | ···················· | NNN················· | 1 |
| Y | 6417 hCoV-19/Wales/PHWC-4950DF/2020\|EPI_ISL_726299\|2020 | ···················· | ···················· | ···················· | ···················· | ···················· | ···················· | |
| Y | 6418 hCoV-19/Wales/PHWC-49A69C/2020\|EPI_ISL_727580\|2020 | ···················· | ···················· | ···················· | ···················· | ···················· | ···················· | |
| Y | 6419 hCoV-19/Wales/PHWC-49A74E/2020\|EPI_ISL_727590\|2020 | ···················· | ···················· | ···················· | ···················· | ···················· | ···················· | |
| Y | 6420 hCoV-19/Wales/PHWC-49A75D/2020\|EPI_ISL_727591\|2020 | ···················· | ···················· | ···················· | ···················· | ···················· | ···················· | |
| Y | 6421 hCoV-19/Wales/PHWC-49A76C/2020\|EPI_ISL_727592\|2020 | ···················· | ···················· | ···················· | ···················· | ···················· | ···················· | |
| Y | 6422 hCoV-19/Wales/PHWC-49A77B/2020\|EPI_ISL_727593\|2020 | ···················· | ···················· | ···················· | ···················· | ···················· | ···················· | |
| Y | 6423 hCoV-19/Wales/PHWC-49A78A/2020\|EPI_ISL_727594\|2020 | ···················· | ······A············· | ···················· | ···················· | ···················· | ···················· | |
| Y | 6424 hCoV-19/Wales/PHWC-49A799/2020\|EPI_ISL_727595\|2020 | ···················· | ···················· | ···················· | ···················· | ···················· | ···················· | |
| Y | 6426 hCoV-19/Wales/PHWC-49A7B7/2020\|EPI_ISL_727597\|2020 | ···················· | ···················· | ···················· | ···················· | ···················· | ···················· | |
| Y | 6427 hCoV-19/Wales/PHWC-49A7D5/2020\|EPI_ISL_727599\|2020 | ···················· | ···················· | ···················· | ···················· | ···················· | ···················· | |
| Y | 6428 hCoV-19/Wales/PHWC-49A7E4/2020\|EPI_ISL_727600\|2020 | ···················· | ···················· | ···················· | ···················· | ···················· | ···················· | |
| Y | 6429 hCoV-19/Wales/PHWC-49A7F3/2020\|EPI_ISL_727601\|2020 | ···················· | ···················· | ···················· | ···················· | ···················· | ···················· | |
| Y | 6430 hCoV-19/Wales/PHWC-49A80F/2020\|EPI_ISL_727602\|2020 | ···················· | ···················· | ···················· | ···················· | ···················· | ···················· | |
| Y | 6432 hCoV-19/Wales/PHWC-49A82D/2020\|EPI_ISL_727604\|2020 | ···················· | ···················· | ···················· | ···················· | ···················· | ···················· | |
| Y | 6433 hCoV-19/Wales/PHWC-49A83C/2020\|EPI_ISL_727605\|2020 | ···················· | ···················· | ···················· | ···················· | ···················· | ···················· | |

| | | | | | | | | |
|---|---|---|---|---|---|---|---|---|
| Y | 6434 | hCoV-19/Wales/PHWC-49A84B/2020\|EPI_ISL_727606\|2020 | ···················· | ···················· | ···················· | ···················· | ···················· | ···················· |
| Y | 6435 | hCoV-19/Wales/PHWC-49A85A/2020\|EPI_ISL_727607\|2020 | ···················· | ···················· | ···················· | ···················· | ···················· | ···················· |
| Y | 6436 | hCoV-19/Wales/PHWC-49A887/2020\|EPI_ISL_727610\|2020 | ···················· | ···················· | ···················· | ···················· | ···················· | ···················· |
| Y | 6437 | hCoV-19/Wales/PHWC-49A896/2020\|EPI_ISL_727611\|2020 | ···················· | ···················· | ···················· | ···················· | ···················· | ···················· |
| Y | 6438 | hCoV-19/Wales/PHWC-49A8B4/2020\|EPI_ISL_727612\|2020 | ···················· | ···················· | ···················· | ···················· | ···················· | ···················· |
| Y | 6439 | hCoV-19/Wales/PHWC-49A8C3/2020\|EPI_ISL_727613\|2020 | ···················· | ···················· | ···················· | ···················· | ···················· | ···················· |
| Y | 6440 | hCoV-19/Wales/PHWC-49A8E1/2020\|EPI_ISL_727615\|2020 | ···················· | ···················· | ···················· | ···················· | ···················· | ···················· |
| Y | 6441 | hCoV-19/Wales/PHWC-49A8F0/2020\|EPI_ISL_727616\|2020 | ···················· | ···················· | ···················· | ···················· | ···················· | ···················· |
| Y | 6442 | hCoV-19/Wales/PHWC-49A91B/2020\|EPI_ISL_727618\|2020 | ···················· | ···················· | ···················· | ···················· | ···················· | ···················· |
| Y | 6443 | hCoV-19/Portugal/PT1885/2020\|EPI_ISL_732100\|2020-1 | ···················· | ···················· | ···················· | ···················· | ···················· | ···················· |
| Y | 6444 | hCoV-19/Portugal/PT1886/2020\|EPI_ISL_732101\|2020-1 | ···················· | ···················· | ···················· | ···················· | ···················· | ···················· |
| Y | 6445 | hCoV-19/Portugal/PT1887/2020\|EPI_ISL_732102\|2020-1 | ···················· | ···················· | ···················· | ···················· | ···················· | ···················· |
| Y | 6446 | hCoV-19/Portugal/PT1888/2020\|EPI_ISL_732103\|2020-1 | ···················· | ···················· | ···················· | ···················· | ···················· | ···················· |
| Y | 6447 | hCoV-19/Portugal/PT1889/2020\|EPI_ISL_732104\|2020-1 | ···················· | ···················· | ···················· | ···················· | ···················· | ···················· |
| Y | 6448 | hCoV-19/Portugal/PT1890/2020\|EPI_ISL_732105\|2020-1 | ···················· | ···················· | ···················· | ···················· | ···················· | ···················· |
| Y | 6449 | hCoV-19/Portugal/PT1891/2020\|EPI_ISL_732106\|2020-1 | ···················· | ···················· | ···················· | ···················· | ···················· | ···················· |
| Y | 6450 | hCoV-19/Portugal/PT1892/2020\|EPI_ISL_732107\|2020-1 | ···················· | ···················· | ···················· | ···················· | ···················· | ···················· |
| Y | 6451 | hCoV-19/Portugal/PT1893/2020\|EPI_ISL_732108\|2020-1 | ···················· | ···················· | ···················· | ···················· | ···················· | ···················· |
| Y | 6452 | hCoV-19/Portugal/PT1894/2020\|EPI_ISL_732109\|2020-1 | ···················· | ···················· | ···················· | ···················· | ···················· | ···················· |
| Y | 6453 | hCoV-19/Portugal/PT1895/2020\|EPI_ISL_732110\|2020-1 | ···················· | ···················· | ···················· | ···················· | ···················· | ···················· |
| Y | 6454 | hCoV-19/Portugal/PT1897/2020\|EPI_ISL_732111\|2020-1 | ···················· | ···················· | ···················· | ···················· | ···················· | ···················· |
| Y | 6455 | hCoV-19/Portugal/PT1898/2020\|EPI_ISL_732112\|2020-1 | ···················· | ···················· | ···················· | ···················· | ···················· | ···················· |
| Y | 6456 | hCoV-19/Portugal/PT1899/2020\|EPI_ISL_732113\|2020-1 | ···················· | ···················· | ···················· | ···················· | ···················· | ···················· |
| Y | 6457 | hCoV-19/Portugal/PT1900/2020\|EPI_ISL_732114\|2020-1 | ···················· | ···················· | ···················· | ···················· | ···················· | ···················· |
| Y | 6458 | hCoV-19/Portugal/PT1901/2020\|EPI_ISL_732115\|2020-1 | ···················· | ···················· | ···················· | ···················· | ···················· | ···················· |
| Y | 6459 | hCoV-19/Portugal/PT1902/2020\|EPI_ISL_732116\|2020-1 | ···················· | ···················· | ···················· | ···················· | ···················· | ···················· |
| Y | 6460 | hCoV-19/Portugal/PT1903/2020\|EPI_ISL_732117\|2020-1 | ···················· | ···················· | ···················· | ···················· | ···················· | ···················· |
| Y | 6461 | hCoV-19/Portugal/PT1904/2020\|EPI_ISL_732118\|2020-1 | ···················· | ···················· | ···················· | ···················· | ···················· | ···················· |
| Y | 6463 | hCoV-19/Portugal/PT1907/2020\|EPI_ISL_732120\|2020-1 | ···················· | ···················· | ···················· | ···················· | ···················· | ···················· |
| Y | 6464 | hCoV-19/Portugal/PT1908/2020\|EPI_ISL_732121\|2020-1 | ···················· | ···················· | ···················· | ···················· | ···················· | ···················· |
| Y | 6465 | hCoV-19/Portugal/PT1909/2020\|EPI_ISL_732122\|2020-1 | ···················· | ···················· | ···················· | ···················· | ···················· | ···················· |
| Y | 6466 | hCoV-19/Portugal/PT1910/2020\|EPI_ISL_732123\|2020-1 | ···················· | ···················· | ···················· | ···················· | ···················· | ···················· |
| Y | 6467 | hCoV-19/Portugal/PT1911/2020\|EPI_ISL_732124\|2020-1 | ···················· | ···················· | ···················· | ···················· | ···················· | ···················· |
| Y | 6468 | hCoV-19/Portugal/PT1912/2020\|EPI_ISL_732125\|2020-1 | ···················· | ···················· | ···················· | ···················· | ···················· | ···················· |
| Y | 6469 | hCoV-19/Portugal/PT1913/2020\|EPI_ISL_732126\|2020-1 | ···················· | ···················· | ···················· | ···················· | ···················· | ···················· |
| Y | 6470 | hCoV-19/Portugal/PT1915/2020\|EPI_ISL_732127\|2020-1 | ···················· | ···················· | ···················· | ···················· | ···················· | ···················· |
| Y | 6471 | hCoV-19/Portugal/PT1916/2020\|EPI_ISL_732128\|2020-1 | ···················· | ···················· | ···················· | ···················· | ···················· | ···················· |
| Y | 6472 | hCoV-19/Portugal/PT1917/2020\|EPI_ISL_732129\|2020-1 | ···················· | ···················· | ···················· | ···················· | ···················· | ···················· |
| Y | 6473 | hCoV-19/Portugal/PT1918/2020\|EPI_ISL_732130\|2020-1 | ···················· | ···················· | ···················· | ···················· | ···················· | ···················· |
| Y | 6474 | hCoV-19/Portugal/PT1919/2020\|EPI_ISL_732131\|2020-1 | ···················· | ···················· | ···················· | ···················· | ···················· | ···················· |
| Y | 6475 | hCoV-19/Portugal/PT1920/2020\|EPI_ISL_732132\|2020-1 | ···················· | ···················· | ···················· | ···················· | ···················· | ···················· |
| Y | 6476 | hCoV-19/Portugal/PT1921/2020\|EPI_ISL_732133\|2020-1 | ···················· | ···················· | ···················· | ···················· | ···················· | ···················· |
| Y | 6477 | hCoV-19/Portugal/PT1922/2020\|EPI_ISL_732134\|2020-1 | ···················· | ···················· | ···················· | ···················· | ···················· | ···················· |
| Y | 6479 | hCoV-19/Portugal/PT1925/2020\|EPI_ISL_732136\|2020-1 | ···················· | ···················· | ···················· | ···················· | ··N················· | ···················· |
| Y | 6480 | hCoV-19/Portugal/PT1926/2020\|EPI_ISL_732137\|2020-1 | ···················· | ···················· | ···················· | ···················· | ···················· | ···················· |
| Y | 6481 | hCoV-19/Portugal/PT1927/2020\|EPI_ISL_732138\|2020-1 | ···················· | ···················· | ···················· | ···················· | ···················· | ···················· |
| Y | 6482 | hCoV-19/Portugal/PT1928/2020\|EPI_ISL_732139\|2020-1 | ···················· | ···················· | ···················· | ···················· | ···················· | ···················· |
| Y | 6483 | hCoV-19/Portugal/PT1929/2020\|EPI_ISL_732140\|2020-1 | ···················· | ···················· | ···················· | ···················· | ···················· | ···················· |
| Y | 6484 | hCoV-19/Portugal/PT1930/2020\|EPI_ISL_732141\|2020-1 | ···················· | ···················· | ···················· | ···················· | ··N················· | ···················· |
| Y | 6485 | hCoV-19/Portugal/PT1931/2020\|EPI_ISL_732142\|2020-1 | ···················· | ···················· | ···················· | ···················· | ···················· | ···················· |
| Y | 6486 | hCoV-19/Portugal/PT1933/2020\|EPI_ISL_732143\|2020-1 | ···················· | ···················· | ···················· | ···················· | ···················· | ···················· |
| Y | 6487 | hCoV-19/Portugal/PT1934/2020\|EPI_ISL_732144\|2020-1 | ···················· | ···················· | ···················· | ···················· | ···················· | ···················· |
| Y | 6488 | hCoV-19/Portugal/PT1935/2020\|EPI_ISL_732145\|2020-1 | ···················· | ···················· | ···················· | ···················· | ···················· | ···················· |
| Y | 6489 | hCoV-19/Portugal/PT1936/2020\|EPI_ISL_732146\|2020-1 | ···················· | ···················· | ···················· | ···················· | ···················· | ···················· |
| Y | 6490 | hCoV-19/Portugal/PT1937/2020\|EPI_ISL_732147\|2020-1 | ···················· | ···················· | ···················· | ···················· | ···················· | ···················· |
| Y | 6491 | hCoV-19/Portugal/PT1681/2020\|EPI_ISL_732148\|2020-1 | ···················· | ···················· | ···················· | ···················· | ···················· | ···················· |
| Y | 6492 | hCoV-19/Portugal/PT1682/2020\|EPI_ISL_732149\|2020-1 | ···················· | ···················· | ···················· | ···················· | ···················· | ···················· |
| Y | 6493 | hCoV-19/Portugal/PT1683/2020\|EPI_ISL_732150\|2020-1 | ···················· | ···················· | ···················· | ···················· | ···················· | ···················· |
| Y | 6494 | hCoV-19/Portugal/PT1684/2020\|EPI_ISL_732151\|2020-1 | ···················· | ···················· | ···················· | ···················· | ···················· | ···················· |
| Y | 6495 | hCoV-19/Portugal/PT1685/2020\|EPI_ISL_732152\|2020-1 | ···················· | ···················· | ···················· | ···················· | ···················· | ···················· |
| Y | 6496 | hCoV-19/Portugal/PT1686/2020\|EPI_ISL_732153\|2020-1 | ···················· | ···················· | ···················· | ···················· | ···················· | ···················· |
| Y | 6497 | hCoV-19/Portugal/PT1687/2020\|EPI_ISL_732154\|2020-1 | ···················· | ···················· | ···················· | ···················· | ···················· | ···················· |
| Y | 6498 | hCoV-19/Portugal/PT1688/2020\|EPI_ISL_732155\|2020-1 | ···················· | ···················· | ···················· | ···················· | ···················· | ···················· |
| Y | 6499 | hCoV-19/Portugal/PT1689/2020\|EPI_ISL_732156\|2020-1 | ···················· | ···················· | ···················· | ···················· | ···················· | ···················· |
| Y | 6500 | hCoV-19/Portugal/PT1690/2020\|EPI_ISL_732157\|2020-1 | ···················· | ···················· | ···················· | ···················· | ···················· | ···················· |
| Y | 6501 | hCoV-19/Portugal/PT1691/2020\|EPI_ISL_732158\|2020-1 | ···················· | ···················· | ···················· | ···················· | ···················· | ···················· |
| Y | 6502 | hCoV-19/Portugal/PT1692/2020\|EPI_ISL_732159\|2020-1 | ···················· | ···················· | ···················· | ···················· | ···················· | ···················· |
| Y | 6503 | hCoV-19/Portugal/PT1693/2020\|EPI_ISL_732160\|2020-1 | ···················· | ···················· | ···················· | ···················· | ···················· | ···················· |
| Y | 6504 | hCoV-19/Portugal/PT1694/2020\|EPI_ISL_732161\|2020-1 | ···················· | ···················· | ···················· | ···················· | ···················· | ···················· |
| Y | 6505 | hCoV-19/Portugal/PT1695/2020\|EPI_ISL_732162\|2020-1 | ···················· | ···················· | ···················· | ···················· | ···················· | ···················· |
| Y | 6506 | hCoV-19/Portugal/PT1696/2020\|EPI_ISL_732163\|2020-1 | ···················· | ···················· | ···················· | ···················· | ···················· | ···················· |
| Y | 6507 | hCoV-19/Portugal/PT1697/2020\|EPI_ISL_732164\|2020-1 | ···················· | ···················· | ···················· | ···················· | ···················· | ···················· |
| Y | 6508 | hCoV-19/Portugal/PT1698/2020\|EPI_ISL_732165\|2020-1 | ···················· | ···················· | ···················· | ···················· | ···················· | ···················· |
| Y | 6509 | hCoV-19/Portugal/PT1699/2020\|EPI_ISL_732166\|2020-1 | ···················· | ···················· | ···················· | ···················· | ···················· | ···················· |
| Y | 6510 | hCoV-19/Portugal/PT1939/2020\|EPI_ISL_732167\|2020-1 | ···················· | ···················· | ···················· | ···················· | ···················· | ···················· |
| Y | 6511 | hCoV-19/Portugal/PT1940/2020\|EPI_ISL_732168\|2020-1 | ···················· | ···················· | ···················· | ···················· | ···················· | ···················· |
| Y | 6512 | hCoV-19/Portugal/PT1941/2020\|EPI_ISL_732169\|2020-1 | ···················· | ···················· | ···················· | ···················· | ···················· | ···················· |
| Y | 6513 | hCoV-19/Portugal/PT1966/2020\|EPI_ISL_732191\|2020-1 | ···················· | ···················· | ···················· | ···················· | ···················· | ···················· |
| Y | 6514 | hCoV-19/Portugal/PT1967/2020\|EPI_ISL_732192\|2020-1 | ···················· | ···················· | ···················· | ···················· | ···················· | ···················· |
| Y | 6515 | hCoV-19/Portugal/PT1968/2020\|EPI_ISL_732193\|2020-1 | ···················· | ···················· | ···················· | ···················· | ···················· | ···················· |

| | | | | | | | | |
|---|---|---|---|---|---|---|---|---|
| Y | 6517 | hCoV-19/Portugal/PT1976/2020\|EPI_ISL_732199\|2020-1 | ···················· | ···················· | ···················· | ···················· | ···················· | ···················· |
| Y | 6518 | hCoV-19/Russia/SVE-RII-MH9000S/2020\|EPI_ISL_733406 | ················G··· | ···················· | ···················· | ···················· | ···················· | ···················· |
| Y | 6520 | hCoV-19/Russia/SVE-RII-MH9113S/2020\|EPI_ISL_733412 | T··················· | ···················· | ···················· | ···················· | ···················· | ···················· |
| Y | 6521 | hCoV-19/Russia/SVE-RII-MH9206S/2020\|EPI_ISL_733414 | ···················· | ···················· | ···················· | ···················· | ···················· | ···················· |
| Y | 6522 | hCoV-19/Russia/KHM-RII-MH9266S/2020\|EPI_ISL_733424 | ···················· | ···················· | ···················· | ···················· | ···················· | ···················· |
| Y | 6523 | hCoV-19/Russia/KDA-RII-MH9294S/2020\|EPI_ISL_733436 | ···················· | ···················· | ···················· | ···················· | ···················· | ···················· |
| Y | 6524 | hCoV-19/Russia/MOS-RII-MH9309S/2020\|EPI_ISL_733439 | ···················· | ···················· | ···················· | ···················· | ···················· | ···················· |
| Y | 6525 | hCoV-19/Russia/MOS-RII-MH9369S/2020\|EPI_ISL_733455 | ···················· | ···················· | ···················· | ···················· | ······T····C········ | ···················· |
| Y | 6526 | hCoV-19/Russia/MOS-RII-MH9370S/2020\|EPI_ISL_733456 | ···················· | ···················· | ···················· | ···················· | ···················· | ···················· |
| Y | 6527 | hCoV-19/Russia/AMU-RII-MH10170S/2020\|EPI_ISL_73346 | ···················· | ···················· | ···················· | ···················· | ···················· | ···················· |
| Y | 6528 | hCoV-19/Russia/AMU-RII-MH10232S/2020\|EPI_ISL_73347 | ···················· | ···················· | ···················· | ···················· | ···················· | ···················· |
| Y | 6529 | hCoV-19/Russia/OMS-ORINFI-MH10275S/2020\|EPI_ISL_73 | ···················· | ···················· | ···················· | ···················· | ···················· | ···················· |
| Y | 6530 | hCoV-19/Russia/OMS-ORINFI-MH10280S/2020\|EPI_ISL_73 | ···················· | ···················· | ···················· | ···················· | ···················· | ···················· |
| Y | 6531 | hCoV-19/Russia/OMS-ORINFI-MH10285M/2020\|EPI_ISL_73 | ···················· | ···················· | ···················· | ···················· | ···················· | ···················· |
| Y | 6532 | hCoV-19/Russia/OMS-ORINFI-MH10288M/2020\|EPI_ISL_73 | ···················· | ···················· | ···················· | ···················· | ···················· | ···················· |
| Y | 6533 | hCoV-19/Russia/OMS-ORINFI-MH10294S/2020\|EPI_ISL_73 | ···················· | ···················· | ···················· | ···················· | ···················· | ···················· |
| Y | 6534 | hCoV-19/Switzerland/ZH-UZH-IMV126/2020\|EPI_ISL_733 | ···················· | ···················· | ···················· | ···················· | ···················· | ···················· |
| Y | 6535 | hCoV-19/Switzerland/ZH-UZH-IMV127/2020\|EPI_ISL_733 | ···················· | ···················· | ···················· | ···················· | ···················· | ···················· |
| Y | 6536 | hCoV-19/Tunisia/63686/2020\|EPI_ISL_733499\|2020-12- | ···················· | ···················· | ···················· | ···················· | ···················· | ···················· |
| Y | 6537 | hCoV-19/Belgium/rega-0502912/2020\|EPI_ISL_734700\|2 | ···················· | ···················· | ···················· | ···················· | ···················· | ···················· |
| Y | 6538 | hCoV-19/Belgium/rega-0503641/2020\|EPI_ISL_734701\|2 | ···················· | ···················· | ···················· | ···················· | ···················· | ···················· |
| Y | 6539 | hCoV-19/Belgium/rega-0503642/2020\|EPI_ISL_734702\|2 | ···················· | ···················· | ···················· | ···················· | ···················· | ···················· |
| Y | 6540 | hCoV-19/Belgium/rega-0503667/2020\|EPI_ISL_734703\|2 | ···················· | ···················· | ···················· | ···················· | ···················· | ···················· |
| Y | 6541 | hCoV-19/Belgium/rega-0505643/2020\|EPI_ISL_734704\|2 | ···················· | ···················· | ···················· | ···················· | ···················· | ···················· |
| Y | 6542 | hCoV-19/Belgium/rega-0506644/2020\|EPI_ISL_734705\|2 | ···················· | ···················· | ···················· | ···················· | ···················· | ···················· |
| Y | 6543 | hCoV-19/Belgium/rega-0506645/2020\|EPI_ISL_734706\|2 | ···················· | ···················· | ···················· | ···················· | ···················· | ···················· |
| Y | 6544 | hCoV-19/Belgium/rega-0507537/2020\|EPI_ISL_734707\|2 | ···················· | ···················· | ···················· | ···················· | ···················· | ···················· |
| Y | 6545 | hCoV-19/Belgium/rega-0507538/2020\|EPI_ISL_734708\|2 | ···················· | ···················· | ···················· | ···················· | ···················· | ···················· |
| Y | 6546 | hCoV-19/Belgium/rega-0507539/2020\|EPI_ISL_734709\|2 | ···················· | ···················· | ···················· | ···················· | ···················· | ···················· |
| Y | 6547 | hCoV-19/Belgium/rega-0508540/2020\|EPI_ISL_734710\|2 | ···················· | ···················· | ···················· | ···················· | ···················· | ···················· |
| Y | 6548 | hCoV-19/Belgium/rega-0508541/2020\|EPI_ISL_734711\|2 | ···················· | ···················· | ···················· | ···················· | ···················· | ···················· |
| Y | 6549 | hCoV-19/Belgium/rega-0508542/2020\|EPI_ISL_734712\|2 | ···················· | ···················· | ···················· | ···················· | ···················· | ···················· |
| Y | 6550 | hCoV-19/Belgium/rega-0508544/2020\|EPI_ISL_734713\|2 | ···················· | ···················· | ···················· | ···················· | ···················· | ···················· |
| Y | 6551 | hCoV-19/Belgium/rega-0508545/2020\|EPI_ISL_734714\|2 | ···················· | ···················· | ···················· | ···················· | ···················· | ···················· |
| Y | 6552 | hCoV-19/Belgium/rega-0508546/2020\|EPI_ISL_734715\|2 | ···················· | ···················· | ···················· | ···················· | ···················· | ···················· |
| Y | 6553 | hCoV-19/Belgium/rega-0508647/2020\|EPI_ISL_734716\|2 | ···················· | ···················· | ···················· | ···················· | ···················· | ···················· |
| Y | 6554 | hCoV-19/Belgium/rega-0508660/2020\|EPI_ISL_734717\|2 | ···················· | ···················· | ···················· | ···················· | ···················· | ···················· |
| Y | 6555 | hCoV-19/Belgium/rega-0509646/2020\|EPI_ISL_734718\|2 | ···················· | ···················· | ···················· | ···················· | ···················· | ···················· |
| Y | 6556 | hCoV-19/Belgium/rega-0509648/2020\|EPI_ISL_734719\|2 | ···················· | ···················· | ···················· | ···················· | ···················· | ···················· |
| Y | 6557 | hCoV-19/Belgium/rega-0509649/2020\|EPI_ISL_734720\|2 | ···················· | ···················· | ···················· | ···················· | ···················· | ···················· |
| Y | 6558 | hCoV-19/Belgium/rega-0509650/2020\|EPI_ISL_734721\|2 | ···················· | ···················· | ···················· | ···················· | ···················· | ···················· |
| Y | 6559 | hCoV-19/Belgium/rega-0509651/2020\|EPI_ISL_734722\|2 | ···················· | ···················· | ···················· | ···················· | ···················· | ···················· |
| Y | 6560 | hCoV-19/Belgium/rega-0509652/2020\|EPI_ISL_734723\|2 | ···················· | ···················· | ···················· | ···················· | ···················· | ···················· |
| Y | 6561 | hCoV-19/Belgium/rega-0510547/2020\|EPI_ISL_734724\|2 | ···················· | ···················· | ···················· | ···················· | ···················· | ···················· |
| Y | 6562 | hCoV-19/Belgium/rega-0512549/2020\|EPI_ISL_734725\|2 | ···················· | ···················· | ···················· | ···················· | ···················· | ···················· |
| Y | 6563 | hCoV-19/Belgium/rega-0512550/2020\|EPI_ISL_734726\|2 | ···················· | ···················· | ···················· | ···················· | ···················· | ···················· |
| Y | 6564 | hCoV-19/Belgium/rega-0512653/2020\|EPI_ISL_734727\|2 | ···················· | ···················· | ···················· | ···················· | ···················· | ···················· |
| Y | 6565 | hCoV-19/Belgium/rega-0513654/2020\|EPI_ISL_734728\|2 | ···················· | ···················· | ···················· | ···················· | ···················· | ···················· |
| Y | 6566 | hCoV-19/Belgium/rega-0513655/2020\|EPI_ISL_734729\|2 | ···················· | ···················· | ···················· | ···················· | ···················· | ···················· |
| Y | 6567 | hCoV-19/Belgium/rega-0513668/2020\|EPI_ISL_734730\|2 | ···················· | ···················· | ···················· | ···················· | ···················· | ···················· |
| Y | 6568 | hCoV-19/Belgium/rega-0514551/2020\|EPI_ISL_734731\|2 | ···················· | ···················· | ···················· | ···················· | ···················· | ···················· |
| Y | 6569 | hCoV-19/Belgium/rega-0514669/2020\|EPI_ISL_734732\|2 | ···················· | ···················· | ···················· | ···················· | ···················· | ···················· |
| Y | 6570 | hCoV-19/Belgium/rega-0514670/2020\|EPI_ISL_734733\|2 | ···················· | ···················· | ···················· | ···················· | ···················· | ···················· |
| Y | 6571 | hCoV-19/Belgium/rega-0514671/2020\|EPI_ISL_734734\|2 | ···················· | ···················· | ···················· | ···················· | ···················· | ···················· |
| Y | 6572 | hCoV-19/Belgium/rega-0516552/2020\|EPI_ISL_734735\|2 | ···················· | ···················· | ···················· | ···················· | ···················· | ···················· |
| Y | 6573 | hCoV-19/Belgium/rega-0517522/2020\|EPI_ISL_734736\|2 | ···················· | ···················· | ···················· | ···················· | ···················· | ···················· |
| Y | 6574 | hCoV-19/Belgium/rega-0517553/2020\|EPI_ISL_734737\|2 | ···················· | ···················· | ···················· | ···················· | ···················· | ···················· |
| Y | 6575 | hCoV-19/Belgium/rega-0517554/2020\|EPI_ISL_734738\|2 | ···················· | ···················· | ···················· | ···················· | ···················· | ···················· |
| Y | 6576 | hCoV-19/Belgium/rega-0517555/2020\|EPI_ISL_734739\|2 | ···················· | ···················· | ···················· | ···················· | ···················· | ···················· |
| Y | 6577 | hCoV-19/Belgium/rega-0521556/2020\|EPI_ISL_734740\|2 | ···················· | ···················· | ···················· | ···················· | ···················· | ···················· |
| Y | 6578 | hCoV-19/Belgium/rega-0521557/2020\|EPI_ISL_734741\|2 | ···················· | ···················· | ···················· | ···················· | ···················· | ···················· |
| Y | 6579 | hCoV-19/Belgium/rega-0521558/2020\|EPI_ISL_734742\|2 | ···················· | ···················· | ···················· | ···················· | ···················· | ···················· |
| Y | 6580 | hCoV-19/Belgium/rega-0521559/2020\|EPI_ISL_734743\|2 | ···················· | ···················· | ···················· | ···················· | ···················· | ···················· |
| Y | 6581 | hCoV-19/Belgium/rega-0526920/2020\|EPI_ISL_734744\|2 | ···················· | ···················· | ···················· | ···················· | ···················· | ···················· |
| Y | 6582 | hCoV-19/Belgium/rega-0528690/2020\|EPI_ISL_734745\|2 | ···················· | ···················· | ···················· | ···················· | ···················· | ···················· |
| Y | 6583 | hCoV-19/Belgium/rega-0528921/2020\|EPI_ISL_734746\|2 | ···················· | ···················· | ···················· | ···················· | ···················· | ···················· |
| Y | 6584 | hCoV-19/Belgium/rega-0602697/2020\|EPI_ISL_734747\|2 | ···················· | ···················· | ···················· | ···················· | ···················· | ···················· |
| Y | 6585 | hCoV-19/Belgium/rega-0603698/2020\|EPI_ISL_734748\|2 | ···················· | ···················· | ···················· | ···················· | ···················· | ···················· |
| Y | 6586 | hCoV-19/Belgium/rega-0610713/2020\|EPI_ISL_734749\|2 | ···················· | ···················· | ···················· | ···················· | ···················· | ···················· |
| Y | 6587 | hCoV-19/Belgium/rega-0611706/2020\|EPI_ISL_734750\|2 | ···················· | ···················· | ···················· | ···················· | ···················· | ···················· |
| Y | 6588 | hCoV-19/Belgium/rega-0611830/2020\|EPI_ISL_734751\|2 | ···················· | ···················· | ···················· | ···················· | ···················· | ···················· |
| Y | 6589 | hCoV-19/Belgium/rega-0612705/2020\|EPI_ISL_734752\|2 | ···················· | ···················· | ···················· | ···················· | ···················· | ···················· |
| Y | 6590 | hCoV-19/Belgium/rega-0615692/2020\|EPI_ISL_734753\|2 | ···················· | ···················· | ···················· | ···················· | ···················· | ···················· |
| Y | 6591 | hCoV-19/Belgium/rega-0615693/2020\|EPI_ISL_734754\|2 | ···················· | ···················· | ···················· | ···················· | ···················· | ···················· |
| Y | 6592 | hCoV-19/Belgium/rega-0615694/2020\|EPI_ISL_734755\|2 | ···················· | ···················· | ···················· | ···················· | ···················· | ···················· |
| Y | 6593 | hCoV-19/Belgium/rega-0615916/2020\|EPI_ISL_734756\|2 | ···················· | ···················· | ···················· | ···················· | ···················· | ···················· |
| Y | 6594 | hCoV-19/Belgium/rega-0616695/2020\|EPI_ISL_734757\|2 | ···················· | ···················· | ···················· | ···················· | ···················· | ···················· |
| Y | 6595 | hCoV-19/Belgium/rega-0616831/2020\|EPI_ISL_734758\|2 | ···················· | ···················· | ···················· | ···················· | ···················· | ···················· |
| Y | 6596 | hCoV-19/Belgium/rega-0616832/2020\|EPI_ISL_734759\|2 | ···················· | ···················· | ···················· | ···················· | ···················· | ···················· |
| Y | 6597 | hCoV-19/Belgium/rega-0616833/2020\|EPI_ISL_734760\|2 | ···················· | ···················· | ···················· | ···················· | ···················· | ···················· |

| | | | | | | | |
|---|---|---|---|---|---|---|---|
| Y | 6598 hCoV-19/Belgium/rega-0616834/2020\|EPI_ISL_734761\|2 | ···················· | ···················· | ···················· | ···················· | ···················· | ···················· |
| Y | 6599 hCoV-19/Belgium/rega-0616835/2020\|EPI_ISL_734762\|2 | ···················· | ···················· | ···················· | ···················· | ···················· | ···················· |
| Y | 6600 hCoV-19/Belgium/rega-0616914/2020\|EPI_ISL_734763\|2 | ···················· | ···················· | ···················· | ···················· | ···················· | ···················· |
| Y | 6601 hCoV-19/Belgium/rega-0616915/2020\|EPI_ISL_734764\|2 | ···················· | ···················· | ···················· | ···················· | ···················· | ···················· |
| Y | 6602 hCoV-19/Belgium/rega-0617691/2020\|EPI_ISL_734765\|2 | ···················· | ···················· | ···················· | ···················· | ···················· | ···················· |
| Y | 6603 hCoV-19/Belgium/rega-0617836/2020\|EPI_ISL_734766\|2 | ···················· | ···················· | ···················· | ···················· | ···················· | ···················· |
| Y | 6604 hCoV-19/Belgium/rega-0617848/2020\|EPI_ISL_734767\|2 | ···················· | ···················· | ···················· | ···················· | ···················· | ···················· |
| Y | 6605 hCoV-19/Belgium/rega-0617913/2020\|EPI_ISL_734768\|2 | ···················· | ···················· | ···················· | ···················· | ···················· | ···················· |
| Y | 6606 hCoV-19/Belgium/rega-0619852/2020\|EPI_ISL_734769\|2 | ···················· | ···················· | ···················· | ···················· | ···················· | ···················· |
| Y | 6607 hCoV-19/Belgium/rega-0619917/2020\|EPI_ISL_734770\|2 | ···················· | ···················· | ···················· | ···················· | ···················· | ···················· |
| Y | 6608 hCoV-19/Belgium/rega-0620707/2020\|EPI_ISL_734771\|2 | ···················· | ···················· | ···················· | ···················· | ···················· | ···················· |
| Y | 6609 hCoV-19/Belgium/rega-0620710/2020\|EPI_ISL_734772\|2 | ···················· | ···················· | ···················· | ···················· | ···················· | ···················· |
| Y | 6610 hCoV-19/Belgium/rega-0620850/2020\|EPI_ISL_734773\|2 | ···················· | ···················· | ···················· | ···················· | ···················· | ···················· |
| Y | 6611 hCoV-19/Belgium/rega-0620851/2020\|EPI_ISL_734774\|2 | ···················· | ···················· | ···················· | ···················· | ···················· | ···················· |
| Y | 6612 hCoV-19/Belgium/rega-0621709/2020\|EPI_ISL_734775\|2 | ···················· | ···················· | ···················· | ···················· | ···················· | ···················· |
| Y | 6613 hCoV-19/Belgium/rega-0622708/2020\|EPI_ISL_734776\|2 | ···················· | ···················· | ···················· | ···················· | ···················· | ···················· |
| Y | 6614 hCoV-19/Belgium/rega-0623700/2020\|EPI_ISL_734777\|2 | ···················· | ···················· | ···················· | ···················· | ···················· | ···················· |
| Y | 6615 hCoV-19/Belgium/rega-0623701/2020\|EPI_ISL_734778\|2 | ···················· | ···················· | ···················· | ···················· | ···················· | ···················· |
| Y | 6616 hCoV-19/Belgium/rega-0623853/2020\|EPI_ISL_734779\|2 | ···················· | ···················· | ···················· | ···················· | ···················· | ···················· |
| Y | 6617 hCoV-19/Belgium/rega-0626822/2020\|EPI_ISL_734780\|2 | ···················· | ···················· | ···················· | ···················· | ···················· | ···················· |
| Y | 6618 hCoV-19/Belgium/rega-0628823/2020\|EPI_ISL_734781\|2 | ···················· | ···················· | ···················· | ···················· | ···················· | ···················· |
| Y | 6619 hCoV-19/Belgium/rega-0629702/2020\|EPI_ISL_734782\|2 | ···················· | ···················· | ···················· | ···················· | ···················· | ···················· |
| Y | 6620 hCoV-19/Belgium/rega-0629824/2020\|EPI_ISL_734783\|2 | ···················· | ···················· | ···················· | ···················· | ···················· | ···················· |
| Y | 6621 hCoV-19/Belgium/rega-0629825/2020\|EPI_ISL_734784\|2 | ···················· | ···················· | ···················· | ···················· | ···················· | ···················· |
| Y | 6622 hCoV-19/Belgium/rega-0630826/2020\|EPI_ISL_734785\|2 | ···················· | ···················· | ···················· | ···················· | ···················· | ···················· |
| Y | 6623 hCoV-19/Belgium/rega-0630827/2020\|EPI_ISL_734786\|2 | ···················· | ···················· | ···················· | ···················· | ···················· | ···················· |
| Y | 6624 hCoV-19/Belgium/rega-0630828/2020\|EPI_ISL_734787\|2 | ···················· | ···················· | ···················· | ···················· | ···················· | ···················· |
| Y | 6625 hCoV-19/Belgium/rega-0701805/2020\|EPI_ISL_734788\|2 | ···················· | ···················· | ···················· | ···················· | ···················· | ···················· |
| Y | 6626 hCoV-19/Belgium/rega-0702806/2020\|EPI_ISL_734789\|2 | ···················· | ···················· | ···················· | ···················· | ···················· | ···················· |
| Y | 6627 hCoV-19/Belgium/rega-0702807/2020\|EPI_ISL_734790\|2 | ···················· | ···················· | ···················· | ···················· | ···················· | ···················· |
| Y | 6628 hCoV-19/Belgium/rega-0703845/2020\|EPI_ISL_734791\|2 | ···················· | ···················· | ···················· | ···················· | ···················· | ···················· |
| Y | 6629 hCoV-19/Belgium/rega-0706712/2020\|EPI_ISL_734792\|2 | ···················· | ···················· | ···················· | ···················· | ···················· | ···················· |
| Y | 6630 hCoV-19/Belgium/rega-0707688/2020\|EPI_ISL_734793\|2 | ···················· | ···················· | ···················· | ···················· | ···················· | ···················· |
| Y | 6631 hCoV-19/Belgium/rega-0707711/2020\|EPI_ISL_734794\|2 | ···················· | ···················· | ···················· | ···················· | ···················· | ···················· |
| Y | 6632 hCoV-19/Belgium/rega-0710918/2020\|EPI_ISL_734795\|2 | ···················· | ···················· | ···················· | ···················· | ···················· | ···················· |
| Y | 6633 hCoV-19/Belgium/rega-0717699/2020\|EPI_ISL_734796\|2 | ···················· | ···················· | ···················· | ···················· | ···················· | ···················· |
| Y | 6634 hCoV-19/Belgium/rega-0717703/2020\|EPI_ISL_734797\|2 | ···················· | ···················· | ···················· | ···················· | ···················· | ···················· |
| Y | 6635 hCoV-19/Belgium/rega-0717846/2020\|EPI_ISL_734798\|2 | ···················· | ···················· | ···················· | ···················· | ···················· | ···················· |
| Y | 6636 hCoV-19/Belgium/rega-0717847/2020\|EPI_ISL_734799\|2 | ···················· | ···················· | ···················· | ···················· | ···················· | ···················· |
| Y | 6637 hCoV-19/Belgium/rega-09211150/2020\|EPI_ISL_738230\| | ···················· | ···················· | ···················· | ···················· | ···················· | ···················· |
| Y | 6638 hCoV-19/Belgium/rega-09211151/2020\|EPI_ISL_738231\| | ···················· | ···················· | ···················· | ···················· | ···················· | ···················· |
| Y | 6639 hCoV-19/Belgium/rega-09221139/2020\|EPI_ISL_738232\| | ···················· | ···················· | ···················· | ···················· | ···················· | ···················· |
| Y | 6640 hCoV-19/Belgium/rega-09221140/2020\|EPI_ISL_738233\| | ···················· | ···················· | ···················· | ···················· | ···················· | ···················· |
| Y | 6641 hCoV-19/Belgium/rega-09221146/2020\|EPI_ISL_738234\| | ···················· | ···················· | ···················· | ···················· | ···················· | ···················· |
| Y | 6642 hCoV-19/Belgium/rega-09221147/2020\|EPI_ISL_738235\| | ···················· | ···················· | ···················· | ···················· | ···················· | ···················· |
| Y | 6643 hCoV-19/Belgium/rega-09221148/2020\|EPI_ISL_738236\| | ···················· | ···················· | ···················· | ···················· | ···················· | ···················· |
| Y | 6644 hCoV-19/Belgium/rega-09221149/2020\|EPI_ISL_738237\| | ···················· | ···················· | ···················· | ···················· | ···················· | ···················· |
| Y | 6645 hCoV-19/Belgium/rega-09231254/2020\|EPI_ISL_738238\| | ···················· | ···················· | ···················· | ···················· | ···················· | ···················· |
| Y | 6646 hCoV-19/Belgium/rega-09241247/2020\|EPI_ISL_738239\| | ···················· | ···················· | ···················· | ···················· | ···················· | ···················· |
| Y | 6647 hCoV-19/Belgium/rega-10061145/2020\|EPI_ISL_738240\| | ···················· | ···················· | ···················· | ···················· | ···················· | ···················· |
| Y | 6648 hCoV-19/France/HDF-rega-0416679/2020\|EPI_ISL_73824 | ···················· | ···················· | ···················· | ···················· | ···················· | ···················· |
| Y | 6649 hCoV-19/France/HDF-rega-0417684/2020\|EPI_ISL_73824 | ···················· | ···················· | ···················· | ···················· | ···················· | ···················· |
| Y | 6650 hCoV-19/Italy/TUS-C266/2020\|EPI_ISL_738243\|2020-03 | ···················· | ···················· | ···················· | ···················· | ···················· | ···················· |
| Y | 6651 hCoV-19/USA/CT-Yale-584/2020\|EPI_ISL_738291\|2020-0 | ···················· | ···················· | ···················· | ···················· | ···················· | ···················· |
| Y | 6652 hCoV-19/USA/CT-Yale-582/2020\|EPI_ISL_738293\|2020-0 | ···················· | ···················· | ···················· | ···················· | ···················· | ···················· |
| Y | 6653 hCoV-19/USA/CT-Yale-581/2020\|EPI_ISL_738294\|2020-0 | ···················· | ···················· | ···················· | ···················· | ···················· | ···················· |
| Y | 6654 hCoV-19/USA/CT-Yale-580/2020\|EPI_ISL_738295\|2020-0 | ···················· | ···················· | ···················· | ···················· | ···················· | ···················· |
| Y | 6655 hCoV-19/USA/CT-Yale-579/2020\|EPI_ISL_738296\|2020-0 | ···················· | ···················· | ···················· | ···················· | ···················· | ···················· |
| Y | 6656 hCoV-19/USA/CT-Yale-578/2020\|EPI_ISL_738297\|2020-0 | ···················· | ···················· | ···················· | ···················· | ···················· | ···················· |
| Y | 6657 hCoV-19/England/NORT-287EE7/2020\|EPI_ISL_725004\|20 | ···················· | ···················· | ···················· | ···················· | ···················· | ···················· |
| Y | 6658 hCoV-19/England/NORT-287EF6/2020\|EPI_ISL_725005\|20 | ···················· | ···················· | ···················· | ···················· | ···················· | ···················· |
| Y | 6659 hCoV-19/England/NORT-287F02/2020\|EPI_ISL_725006\|20 | ···················· | ···················· | ···················· | ···················· | ···················· | ···················· |
| Y | 6660 hCoV-19/England/NORT-287F7B/2020\|EPI_ISL_725007\|20 | ···················· | ···················· | ···················· | ···················· | ···················· | ···················· |
| Y | 6661 hCoV-19/England/NORT-287FE4/2020\|EPI_ISL_725008\|20 | ···················· | ···················· | ···················· | ···················· | ···················· | ···················· |
| Y | 6662 hCoV-19/England/NORT-28800E/2020\|EPI_ISL_725009\|20 | ···················· | ···················· | ···················· | ···················· | ···················· | ···················· |
| Y | 6663 hCoV-19/England/NORT-2881ED/2020\|EPI_ISL_725011\|20 | ···················· | ···················· | ···················· | ···················· | ···················· | ···················· |
| Y | 6664 hCoV-19/England/NORT-288411/2020\|EPI_ISL_725012\|20 | ···················· | ···················· | ···················· | ···················· | ···················· | ···················· |
| Y | 6665 hCoV-19/England/NORT-28844E/2020\|EPI_ISL_725014\|20 | ···················· | ···················· | ···················· | ···················· | ···················· | ···················· |
| Y | 6666 hCoV-19/England/NORT-28846C/2020\|EPI_ISL_725015\|20 | ···················· | ···················· | ···················· | ···················· | ···················· | ···················· |
| Y | 6667 hCoV-19/England/NORT-2884A8/2020\|EPI_ISL_725016\|20 | ···················· | ···················· | ···················· | ···················· | ···················· | ···················· |
| Y | 6668 hCoV-19/England/NORT-28851E/2020\|EPI_ISL_725020\|20 | ···················· | ···················· | ···················· | ···················· | ·····T·············· | ···················· |
| Y | 6669 hCoV-19/England/NORT-28852D/2020\|EPI_ISL_725021\|20 | ···················· | ···················· | ···················· | ···················· | ·····T·············· | ···················· |
| Y | 6670 hCoV-19/England/NORT-28853C/2020\|EPI_ISL_725022\|20 | ···················· | ···················· | ···················· | ···················· | ···················· | ···················· |
| Y | 6671 hCoV-19/England/NORT-288578/2020\|EPI_ISL_725023\|20 | ···················· | ···················· | ···················· | ···················· | ·····T·············· | ···················· |
| Y | 6672 hCoV-19/England/NORT-2885D2/2020\|EPI_ISL_725025\|20 | ···················· | ···················· | ···················· | ···················· | ···················· | ···················· |
| Y | 6673 hCoV-19/England/NORT-2885E1/2020\|EPI_ISL_725026\|20 | ···················· | ···················· | ···················· | ···················· | ···················· | ···················· |
| Y | 6674 hCoV-19/England/NORT-2885F0/2020\|EPI_ISL_725027\|20 | ···················· | ···················· | ···················· | ···················· | ·····T·············· | ···················· |
| Y | 6676 hCoV-19/England/NORW-21F5D0/2020\|EPI_ISL_725031\|20 | ···················· | ···················· | ···················· | ···················· | ···················· | ···················· |
| Y | 6677 hCoV-19/England/NORW-21F5FE/2020\|EPI_ISL_725033\|20 | ···················· | ···················· | ···················· | ···················· | ···················· | ···················· |
| Y | 6678 hCoV-19/England/NORW-21F646/2020\|EPI_ISL_725036\|20 | ···················· | ·A·················· | ···················· | ···················· | ···················· | ··A················· |

| | | | | | | | | |
|---|---|---|---|---|---|---|---|---|
| Y | 6679 hCoV-19/England/NORW-21F682/2020\|EPI_ISL_725038\|20 | ···················· | ···················· | ···················· | ···················· | ···················· | ···················· | |
| Y | 6680 hCoV-19/England/NORW-21F6A0/2020\|EPI_ISL_725039\|20 | ···················· | ···················· | ···················· | ···················· | ···················· | ···················· | |
| Y | 6683 hCoV-19/Wales/PHWC-495109/2020\|EPI_ISL_726302\|2020 | ···················· | ···················· | ···················· | ···················· | ···················· | ···················· | |
| Y | 6685 hCoV-19/Wales/PHWC-495136/2020\|EPI_ISL_726304\|2020 | ···················· | ···················· | ···················· | ···················· | ···················· | ···················· | |
| Y | 6686 hCoV-19/Wales/PHWC-495154/2020\|EPI_ISL_726306\|2020 | ···················· | ···················· | ···················· | ···················· | ···················· | ···················· | |
| Y | 6687 hCoV-19/Wales/PHWC-495163/2020\|EPI_ISL_726307\|2020 | ···················· | ···················· | ···················· | ···················· | NNN················· | ···················· | 1 |
| Y | 6688 hCoV-19/Wales/PHWC-4951AF/2020\|EPI_ISL_726308\|2020 | ···················· | ···················· | ···················· | ···················· | ···················· | ···················· | |
| Y | 6691 hCoV-19/Wales/PHWC-495206/2020\|EPI_ISL_726312\|2020 | ···················· | ···················· | ···················· | ···················· | ···················· | NN·················· | |
| Y | 6692 hCoV-19/Wales/PHWC-495224/2020\|EPI_ISL_726314\|2020 | ···················· | ···················· | ···················· | ···················· | ···················· | ···················· | |
| Y | 6695 hCoV-19/Wales/PHWC-4952AC/2020\|EPI_ISL_726321\|2020 | ···················· | ···················· | ···················· | ···················· | ···················· | ···················· | |
| Y | 6697 hCoV-19/Wales/PHWC-4952D9/2020\|EPI_ISL_726324\|2020 | ···················· | ···················· | ···················· | ···················· | ···················· | ···················· | |
| Y | 6707 hCoV-19/Wales/PHWC-4953A9/2020\|EPI_ISL_726337\|2020 | ···················· | ···················· | ···················· | ···················· | ···················· | ···················· | |
| Y | 6711 hCoV-19/Wales/PHWC-4953F4/2020\|EPI_ISL_726342\|2020 | ···················· | ···················· | ···················· | ···················· | ···················· | ···················· | |
| Y | 6715 hCoV-19/Wales/PHWC-49545B/2020\|EPI_ISL_726348\|2020 | ···················· | ···················· | ···················· | ···················· | ···················· | ···················· | |
| Y | 6716 hCoV-19/Wales/PHWC-49546A/2020\|EPI_ISL_726349\|2020 | ···················· | ···················· | ···················· | ···················· | ···················· | ···················· | |
| Y | 6717 hCoV-19/Wales/PHWC-495479/2020\|EPI_ISL_726350\|2020 | ···················· | ···················· | ···················· | ···················· | ···················· | ···················· | |
| Y | 6719 hCoV-19/Wales/PHWC-4954A6/2020\|EPI_ISL_726352\|2020 | ···················· | ···················· | ···················· | ···················· | ···················· | ···················· | |
| Y | 6720 hCoV-19/Wales/PHWC-4954B5/2020\|EPI_ISL_726353\|2020 | ···················· | ···················· | ···················· | ···················· | ···················· | ···················· | |
| Y | 6722 hCoV-19/Wales/PHWC-4954D3/2020\|EPI_ISL_726355\|2020 | ···················· | ···················· | ···················· | ···················· | ···················· | ···················· | |
| Y | 6723 hCoV-19/Wales/PHWC-4954E2/2020\|EPI_ISL_726356\|2020 | ···················· | ···················· | ···················· | ···················· | ···················· | ···················· | |
| Y | 6728 hCoV-19/Wales/PHWC-495576/2020\|EPI_ISL_726363\|2020 | ···················· | ···················· | ···················· | ···················· | ···················· | ···················· | |
| Y | 6729 hCoV-19/Wales/PHWC-495594/2020\|EPI_ISL_726365\|2020 | ···················· | ···················· | ···················· | ···················· | ···················· | ···················· | |
| Y | 6730 hCoV-19/Wales/PHWC-4955A3/2020\|EPI_ISL_726366\|2020 | ···················· | ···················· | ···················· | ···················· | ···················· | ···················· | |
| Y | 6734 hCoV-19/Wales/PHWC-49A869/2020\|EPI_ISL_727608\|2020 | ···················· | ···················· | ···················· | ···················· | ···················· | ···················· | |
| Y | 6735 hCoV-19/Wales/PHWC-49A878/2020\|EPI_ISL_727609\|2020 | ···················· | ···················· | ···················· | ···················· | ···················· | ···················· | |
| Y | 6736 hCoV-19/Wales/PHWC-49A948/2020\|EPI_ISL_727620\|2020 | ···················· | ···················· | ···················· | ···················· | ···················· | ···················· | |
| Y | 6737 hCoV-19/Wales/PHWC-49A957/2020\|EPI_ISL_727621\|2020 | ···················· | ···················· | ···················· | ···················· | ···················· | ···················· | |
| Y | 6738 hCoV-19/Wales/PHWC-49A966/2020\|EPI_ISL_727622\|2020 | ···················· | ···················· | ···················· | ···················· | ···················· | ···················· | |
| Y | 6740 hCoV-19/Wales/PHWC-49A993/2020\|EPI_ISL_727624\|2020 | ···················· | ···················· | ···················· | ···················· | ···················· | ···················· | |
| Y | 6741 hCoV-19/Wales/PHWC-49A9C0/2020\|EPI_ISL_727627\|2020 | ···················· | ···················· | ···················· | ···················· | ···················· | ···················· | |
| Y | 6743 hCoV-19/Wales/PHWC-49A9EE/2020\|EPI_ISL_727629\|2020 | ···················· | ···················· | ···················· | ···················· | ···················· | ···················· | |
| Y | 6744 hCoV-19/Wales/PHWC-49A9FD/2020\|EPI_ISL_727630\|2020 | ···················· | ···················· | ···················· | ···················· | ···················· | ···················· | |
| Y | 6745 hCoV-19/Wales/PHWC-49AA09/2020\|EPI_ISL_727631\|2020 | ···················· | ···················· | ···················· | ···················· | ···················· | ···················· | |
| Y | 6747 hCoV-19/Wales/PHWC-49AA27/2020\|EPI_ISL_727633\|2020 | ···················· | ···················· | ···················· | ···················· | ···················· | ···················· | |
| Y | 6748 hCoV-19/Wales/PHWC-49AA36/2020\|EPI_ISL_727634\|2020 | ···················· | ···················· | ···················· | ···················· | ···················· | ···················· | |
| Y | 6749 hCoV-19/Wales/PHWC-49AA45/2020\|EPI_ISL_727635\|2020 | ···················· | ···················· | ···················· | ···················· | ···················· | ···················· | |
| Y | 6750 hCoV-19/Wales/PHWC-49AA54/2020\|EPI_ISL_727636\|2020 | ···················· | ···················· | ···················· | ···················· | ···················· | ···················· | |
| Y | 6751 hCoV-19/Wales/PHWC-49AA63/2020\|EPI_ISL_727637\|2020 | ···················· | ···················· | ···················· | ···················· | ···················· | ···················· | |
| Y | 6754 hCoV-19/Wales/PHWC-49AABE/2020\|EPI_ISL_727642\|2020 | ···················· | ···················· | ···················· | ···················· | ···················· | ···················· | |
| Y | 6755 hCoV-19/Wales/PHWC-49AACD/2020\|EPI_ISL_727643\|2020 | ···················· | ···················· | ···················· | ···················· | ···················· | ···················· | |
| Y | 6756 hCoV-19/Wales/PHWC-49AADC/2020\|EPI_ISL_727644\|2020 | ···················· | ···················· | ···················· | ···················· | ···················· | ···················· | |
| Y | 6758 hCoV-19/Wales/PHWC-49AB06/2020\|EPI_ISL_727646\|2020 | ···················· | ···················· | ···················· | ···················· | ···················· | ···················· | |
| Y | 6759 hCoV-19/Wales/PHWC-49AB15/2020\|EPI_ISL_727647\|2020 | ···················· | ···················· | ···················· | ···················· | ···················· | ···················· | |
| Y | 6760 hCoV-19/Wales/PHWC-49AB33/2020\|EPI_ISL_727649\|2020 | ···················· | ···················· | ···················· | ···················· | ···················· | ···················· | |
| Y | 6761 hCoV-19/Wales/PHWC-49AB42/2020\|EPI_ISL_727650\|2020 | ···················· | ···················· | ···················· | ···················· | ···················· | ···················· | |
| Y | 6762 hCoV-19/Wales/PHWC-49AB51/2020\|EPI_ISL_727651\|2020 | ···················· | ···················· | ···················· | ···················· | ···················· | ···················· | |
| Y | 6763 hCoV-19/Wales/PHWC-49AB60/2020\|EPI_ISL_727652\|2020 | ···················· | ···················· | ···················· | ···················· | ···················· | ···················· | |
| Y | 6764 hCoV-19/Wales/PHWC-49AB7F/2020\|EPI_ISL_727653\|2020 | ···················· | ···················· | ···················· | ···················· | ···················· | ···················· | |
| Y | 6767 hCoV-19/Wales/PHWC-49ABAC/2020\|EPI_ISL_727656\|2020 | ···················· | ···················· | ···················· | ···················· | ···················· | ···················· | |
| Y | 6768 hCoV-19/Wales/PHWC-49ABBB/2020\|EPI_ISL_727657\|2020 | ···················· | ···················· | ···················· | ···················· | ···················· | ···················· | |
| Y | 6772 hCoV-19/Wales/PHWC-49ABF7/2020\|EPI_ISL_727661\|2020 | ···················· | ···················· | ···················· | ···················· | ···················· | ···················· | |
| Y | 6773 hCoV-19/Wales/PHWC-49AC03/2020\|EPI_ISL_727662\|2020 | ···················· | ···················· | ···················· | ···················· | ···················· | ···················· | |
| Y | 6776 hCoV-19/Wales/PHWC-49AC30/2020\|EPI_ISL_727665\|2020 | ···················· | ···················· | ···················· | ···················· | ···················· | ···················· | |
| Y | 6777 hCoV-19/Wales/PHWC-49AC4F/2020\|EPI_ISL_727666\|2020 | ···················· | ···················· | ···················· | ···················· | ···················· | ···················· | |
| Y | 6781 hCoV-19/Wales/PHWC-49ACC7/2020\|EPI_ISL_727673\|2020 | ···················· | ···················· | ···················· | ···················· | ···················· | ···················· | |
| Y | 6782 hCoV-19/Wales/PHWC-49ACD6/2020\|EPI_ISL_727674\|2020 | ···················· | ···················· | ···················· | ···················· | ···················· | ···················· | |
| Y | 6789 hCoV-19/Wales/PHWC-49AD88/2020\|EPI_ISL_727683\|2020 | ···················· | ···················· | ···················· | ···················· | ···················· | ···················· | |
| Y | 6792 hCoV-19/Wales/PHWC-49ADD3/2020\|EPI_ISL_727688\|2020 | ···················· | ···················· | ···················· | ···················· | ···················· | ···················· | |
| Y | 6798 hCoV-19/Wales/PHWC-49AE3A/2020\|EPI_ISL_727694\|2020 | ···················· | ···················· | ···················· | ···················· | NNN················· | ···················· | 1 |
| Y | 6804 hCoV-19/USA/WI-UW-2342/2020\|EPI_ISL_728108\|2020-11 | ···················· | ···················· | ···················· | ···················· | ···················· | ···················· | |
| Y | 6806 hCoV-19/Germany/DE-SH-ChVir9987/2020\|EPI_ISL_72943 | ···················· | ···················· | ···················· | ···················· | ···················· | ···················· | |
| Y | 6807 hCoV-19/Germany/DE-SH-ChVir9992/2020\|EPI_ISL_72943 | ···················· | ···················· | ···················· | ···················· | ···················· | ···················· | |
| Y | 6808 hCoV-19/Germany/DE-SH-ChVir20008/2020\|EPI_ISL_7294 | ···················· | ···················· | ···················· | ···················· | ···················· | ···················· | |
| Y | 6809 hCoV-19/Germany/DE-SH-ChVir20009/2020\|EPI_ISL_7294 | ···················· | ···················· | ···················· | ···················· | ···················· | ···················· | |
| Y | 6810 hCoV-19/Germany/DE-SH-ChVir20013/2020\|EPI_ISL_7294 | ···················· | ···················· | ···················· | ···················· | ···················· | ···················· | |
| Y | 6811 hCoV-19/Germany/DE-SH-ChVir20017/2020\|EPI_ISL_7294 | ···················· | ···················· | ···················· | ···················· | ···················· | ···················· | |
| Y | 6812 hCoV-19/Germany/DE-SH-ChVir20022/2020\|EPI_ISL_7294 | ···················· | ···················· | ···················· | ···················· | ···················· | ···················· | |
| Y | 6813 hCoV-19/Germany/DE-SH-ChVir20027/2020\|EPI_ISL_7294 | ···················· | ···················· | ···················· | ···················· | ···················· | ···················· | |
| Y | 6814 hCoV-19/Germany/DE-HH-ChVir20032/2020\|EPI_ISL_7294 | ···················· | ···················· | ···················· | ···················· | ···················· | ···················· | |
| Y | 6815 hCoV-19/Germany/DE-SH-ChVir20033/2020\|EPI_ISL_7294 | ···················· | ···················· | ···················· | ···················· | ···················· | ···················· | |
| Y | 6816 hCoV-19/Jordan/AM-ALSR-5165/2020\|EPI_ISL_730404\|20 | ···················· | ···················· | ···················· | ···················· | ···················· | ···················· | |
| Y | 6817 hCoV-19/Jordan/AM-ALSR-5166/2020\|EPI_ISL_730405\|20 | ···················· | ·A·················· | ···················· | ···················· | ···················· | ··A················· | |
| Y | 6818 hCoV-19/Jordan/AM-ALSR-5174/2020\|EPI_ISL_730413\|20 | ···················· | ·A·················· | ···················· | ···················· | ···················· | ··A················· | |
| Y | 6819 hCoV-19/Jordan/AM-ALSR-5175/2020\|EPI_ISL_730414\|20 | ···················· | ···················· | ···················· | ···················· | ·····T·············· | ···················· | |
| Y | 6820 hCoV-19/Jordan/AM-ALSR-5176/2020\|EPI_ISL_730415\|20 | ···················· | ···················· | ···················· | ···················· | ·····T·············· | ···················· | |
| Y | 6821 hCoV-19/Jordan/AM-ALSR-5177/2020\|EPI_ISL_730416\|20 | ···················· | ···················· | ···················· | ···················· | ···················· | ···················· | |
| Y | 6822 hCoV-19/Jordan/AM-ALSR-5178/2020\|EPI_ISL_730417\|20 | ···················· | ···················· | ···················· | ···················· | ···················· | ···················· | |
| Y | 6823 hCoV-19/Jordan/AM-ALSR-5179/2020\|EPI_ISL_730418\|20 | ···················· | ···················· | ···················· | ···················· | ···················· | ···················· | |
| Y | 6824 hCoV-19/Jordan/AM-ALSR-5180/2020\|EPI_ISL_730419\|20 | ···················· | ·A·················· | ···················· | ···················· | ···················· | ··A················· | |
| Y | 6825 hCoV-19/Jordan/AM-ALSR-5181/2020\|EPI_ISL_730420\|20 | ···················· | ···················· | ···················· | ···················· | ···················· | ···················· | |

| | | | | | | | |
|---|---|---|---|---|---|---|---|
| Y | 6826 hCoV-19/Jordan/AM-ALSR-5182/2020\|EPI_ISL_730421\|20 | .................... | .................... | ..................... | .................... | .................... | .................... |
| Y | 6827 hCoV-19/Jordan/AM-ALSR-5183/2020\|EPI_ISL_730422\|20 | .................... | .................... | ..................... | .................... | .................... | .................... |
| Y | 6828 hCoV-19/Jordan/AM-ALSR-5184/2020\|EPI_ISL_730423\|20 | .................... | .................... | ..................... | .................... | .................... | .................... |
| Y | 6829 hCoV-19/England/ALDP-C3D332/2020\|EPI_ISL_731735\|20 | .................... | .................... | ..................... | .................... | .................... | .................... |
| Y | 6830 hCoV-19/England/ALDP-C3B282/2020\|EPI_ISL_731736\|20 | .................... | .................... | ..................... | .................... | .................... | .................... |
| Y | 6831 hCoV-19/England/ALDP-C3BB78/2020\|EPI_ISL_731744\|20 | .................... | .................... | ..................... | .................... | .................... | .................... |
| Y | 6832 hCoV-19/England/ALDP-C3BFE5/2020\|EPI_ISL_731745\|20 | .................... | .................... | ..................... | .................... | .................... | .................... |
| Y | 6833 hCoV-19/England/ALDP-C3CAF2/2020\|EPI_ISL_731747\|20 | .................... | .................... | ..................... | .................... | .................... | .................... |
| Y | 6834 hCoV-19/England/ALDP-C3BDFA/2020\|EPI_ISL_731748\|20 | .................... | .................... | ..................... | .................... | .................... | .................... |
| Y | 6835 hCoV-19/England/ALDP-C3CB77/2020\|EPI_ISL_731749\|20 | .................... | .................... | ..................... | .................... | .................... | .................... |
| Y | 6836 hCoV-19/England/ALDP-C3D578/2020\|EPI_ISL_731751\|20 | .................... | .................... | ..................... | .................... | .................... | .................... |
| Y | 6837 hCoV-19/England/ALDP-C3D165/2020\|EPI_ISL_731752\|20 | .................... | .................... | ..................... | .................... | .................... | .................... |
| Y | 6838 hCoV-19/England/ALDP-C3BF21/2020\|EPI_ISL_731753\|20 | .................... | .................... | ..................... | .................... | .................... | .................... |
| Y | 6839 hCoV-19/England/ALDP-C3CBA4/2020\|EPI_ISL_731754\|20 | .................... | .................... | ..................... | .................... | .................... | .................... |
| Y | 6840 hCoV-19/Portugal/PT1942/2020\|EPI_ISL_732170\|2020-1 | .................... | .................... | ..................... | .................... | .................... | .................... |
| Y | 6841 hCoV-19/Portugal/PT1943/2020\|EPI_ISL_732171\|2020-1 | .................... | .................... | ..................... | .................... | .................... | .................... |
| Y | 6842 hCoV-19/Portugal/PT1945/2020\|EPI_ISL_732172\|2020-1 | .................... | .................... | ..................... | .................... | .................... | .................... |
| Y | 6843 hCoV-19/Portugal/PT1946/2020\|EPI_ISL_732173\|2020-1 | .................... | .................... | ..................... | .................... | .................... | .................... |
| Y | 6844 hCoV-19/Portugal/PT1947/2020\|EPI_ISL_732174\|2020-1 | .................... | .................... | ..................... | .................... | .................... | .................... |
| Y | 6845 hCoV-19/Portugal/PT1948/2020\|EPI_ISL_732175\|2020-1 | .................... | .................... | ..................... | .................... | .................... | .................... |
| Y | 6846 hCoV-19/Portugal/PT1949/2020\|EPI_ISL_732176\|2020-1 | .................... | .................... | ..................... | .................... | .................... | .................... |
| Y | 6847 hCoV-19/Portugal/PT1950/2020\|EPI_ISL_732177\|2020-1 | .................... | .................... | ..................... | .................... | .................... | .................... |
| Y | 6848 hCoV-19/Portugal/PT1951/2020\|EPI_ISL_732178\|2020-1 | .................... | .................... | ..................... | .................... | .................... | .................... |
| Y | 6849 hCoV-19/Portugal/PT1952/2020\|EPI_ISL_732179\|2020-1 | .................... | .................... | ..................... | .................... | .................... | .................... |
| Y | 6850 hCoV-19/Portugal/PT1953/2020\|EPI_ISL_732180\|2020-1 | .................... | .................... | ..................... | .................... | .................... | .................... |
| Y | 6851 hCoV-19/Portugal/PT1954/2020\|EPI_ISL_732181\|2020-1 | .................... | .................... | ..................... | .................... | .................... | .................... |
| Y | 6852 hCoV-19/Portugal/PT1955/2020\|EPI_ISL_732182\|2020-1 | .................... | .................... | ..................... | .................... | .................... | .................... |
| Y | 6853 hCoV-19/Portugal/PT1956/2020\|EPI_ISL_732183\|2020-1 | .................... | .................... | ..................... | .................... | .................... | .................... |
| Y | 6854 hCoV-19/Portugal/PT1957/2020\|EPI_ISL_732184\|2020-1 | .................... | .................... | ..................... | .................... | .................... | .................... |
| Y | 6855 hCoV-19/Portugal/PT1958/2020\|EPI_ISL_732185\|2020-1 | .................... | .................... | ..................... | .................... | .................... | .................... |
| Y | 6856 hCoV-19/Portugal/PT1959/2020\|EPI_ISL_732186\|2020-1 | .................... | .................... | ..................... | .................... | .................... | .................... |
| Y | 6857 hCoV-19/Portugal/PT1960/2020\|EPI_ISL_732187\|2020-1 | .................... | .................... | ..................... | .................... | .................... | .................... |
| Y | 6858 hCoV-19/Portugal/PT1962/2020\|EPI_ISL_732188\|2020-1 | .................... | .................... | ..................... | .................... | .................... | .................... |
| Y | 6859 hCoV-19/Portugal/PT1964/2020\|EPI_ISL_732189\|2020-1 | .................... | ...A................ | ..................... | .................... | .................... | .................... |
| Y | 6860 hCoV-19/Portugal/PT1965/2020\|EPI_ISL_732190\|2020-1 | .................... | .................... | ..................... | .................... | .................... | .................... |
| Y | 6861 hCoV-19/Portugal/PT1969/2020\|EPI_ISL_732194\|2020-1 | .................... | .................... | ..................... | .................... | .................... | .................... |
| Y | 6862 hCoV-19/Portugal/PT1970/2020\|EPI_ISL_732195\|2020-1 | .................... | .................... | ..................... | .................... | .................... | .................... |
| Y | 6863 hCoV-19/Portugal/PT1972/2020\|EPI_ISL_732196\|2020-1 | .................... | .................... | ..................... | .................... | .................... | .................... |
| Y | 6864 hCoV-19/Portugal/PT1973/2020\|EPI_ISL_732197\|2020-1 | .................... | ...A................ | ..................... | .................... | .................... | .................... |
| Y | 6865 hCoV-19/Czech Republic/NRL_10374/2020\|EPI_ISL_7370 | .................... | .................... | ..................... | .................... | .................... | .................... |
| Y | 6866 hCoV-19/Czech Republic/NRL_10386/2020\|EPI_ISL_7370 | .................... | .................... | ..................... | .................... | .................... | .................... |
| Y | 6867 hCoV-19/Czech Republic/NRL_10398/2020\|EPI_ISL_7370 | .................... | .................... | ..................... | .................... | .................... | .................... |
| Y | 6868 hCoV-19/Czech Republic/NRL_10405/2020\|EPI_ISL_7370 | .................... | .................... | ..................... | .................... | .................... | .................... |
| Y | 6869 hCoV-19/Czech Republic/NRL_10406/2020\|EPI_ISL_7370 | .................... | .................... | ..................... | .................... | .................... | .................... |
| Y | 6870 hCoV-19/Czech Republic/NRL_10410/2020\|EPI_ISL_7370 | .................... | .................... | ..................... | .................... | .................... | .................... |
| Y | 6871 hCoV-19/Czech Republic/NRL_10411/2020\|EPI_ISL_7370 | .................... | .................... | ..................... | .................... | .................... | .................... |
| Y | 6872 hCoV-19/Czech Republic/NRL_10416/2020\|EPI_ISL_7370 | .................... | .................... | ..................... | .................... | .................... | .................... |
| Y | 6873 hCoV-19/Czech Republic/NRL_10417/2020\|EPI_ISL_7370 | .................... | .................... | ..................... | .................... | .................... | .................... |
| Y | 6874 hCoV-19/Czech Republic/NRL_10418/2020\|EPI_ISL_7370 | .................... | .................... | ..................... | .................... | .................... | .................... |
| Y | 6875 hCoV-19/Czech Republic/NRL_10419/2020\|EPI_ISL_7370 | .................... | .................... | ..................... | .................... | .................... | .................... |
| Y | 6876 hCoV-19/Czech Republic/NRL_10420/2020\|EPI_ISL_7370 | .................... | .................... | ..................... | .................... | .................... | .................... |
| Y | 6877 hCoV-19/Czech Republic/NRL_10421/2020\|EPI_ISL_7370 | .................... | .................... | ..................... | .................... | .................... | .................... |
| Y | 6878 hCoV-19/Czech Republic/NRL_10472/2020\|EPI_ISL_7370 | .................... | .................... | ..................... | .................... | .................... | .................... |
| Y | 6879 hCoV-19/Czech Republic/NRL_10749/2020\|EPI_ISL_7370 | .................... | .................... | ..................... | .................... | .................... | .................... |
| Y | 6880 hCoV-19/Czech Republic/NRL_10750/2020\|EPI_ISL_7370 | .................... | .................... | ..................... | .................... | .................... | .................... |
| Y | 6881 hCoV-19/Czech Republic/NRL_10804/2020\|EPI_ISL_7370 | .................... | .................... | ..................... | .................... | .................... | .................... |
| Y | 6882 hCoV-19/Czech Republic/NRL_10875/2020\|EPI_ISL_7370 | .................... | .................... | ..................... | .................... | .................... | .................... |
| Y | 6883 hCoV-19/Czech Republic/NRL_10882/2020\|EPI_ISL_7370 | .................... | .................... | ..................... | .................... | .................... | .................... |
| Y | 6884 hCoV-19/Czech Republic/NRL_10908/2020\|EPI_ISL_7370 | .................... | .................... | ..................... | .................... | .................... | .................... |
| Y | 6885 hCoV-19/USA/CT-Yale-397/2020\|EPI_ISL_738310\|2020-0 | .................... | .................... | ..................... | .................... | .................... | .................... |
| Y | 6886 hCoV-19/USA/CT-Yale-395/2020\|EPI_ISL_738312\|2020-0 | .................... | .................... | ..................... | .................... | .................... | .................... |
| Y | 6887 hCoV-19/Romania/USAFSAM-S542/2020\|EPI_ISL_738326\|2 | .................... | .................... | ..................... | .................... | .................... | .................... |
| Y | 6888 hCoV-19/Romania/USAFSAM-S543/2020\|EPI_ISL_738327\|2 | .................... | .................... | ..................... | .................... | .................... | .................... |
| Y | 6889 hCoV-19/Romania/USAFSAM-S544/2020\|EPI_ISL_738328\|2 | .................... | ...A................ | ..................... | .................... | .................... | .................... |
| Y | 6890 hCoV-19/Romania/USAFSAM-S546/2020\|EPI_ISL_738329\|2 | .................... | .................... | ..................... | .................... | .................... | .................... |
| Y | 6891 hCoV-19/Romania/USAFSAM-S547/2020\|EPI_ISL_738330\|2 | .................... | ...A................ | ..................... | .................... | .................... | .................... |
| Y | 6892 hCoV-19/Romania/USAFSAM-S548/2020\|EPI_ISL_738331\|2 | .................... | ...A................ | ..................... | .................... | .................... | .................... |
| Y | 6893 hCoV-19/Romania/USAFSAM-S549/2020\|EPI_ISL_738332\|2 | .................... | ...A................ | ..................... | .................... | .................... | .................... |
| Y | 6894 hCoV-19/Romania/USAFSAM-S550/2020\|EPI_ISL_738333\|2 | .................... | .................... | ..................... | .................... | .................... | .................... |
| Y | 6895 hCoV-19/Romania/USAFSAM-S552/2020\|EPI_ISL_738334\|2 | .................... | .................... | ..................... | .................... | .................... | .................... |
| Y | 6896 hCoV-19/Romania/USAFSAM-S554/2020\|EPI_ISL_738335\|2 | .................... | ...A................ | ..................... | .................... | .................... | .................... |
| Y | 6897 hCoV-19/Romania/USAFSAM-S555/2020\|EPI_ISL_738336\|2 | .................... | ...A................ | ..................... | .................... | .................... | .................... |
| Y | 6898 hCoV-19/Romania/USAFSAM-S557/2020\|EPI_ISL_738337\|2 | .................... | ...A................ | ..................... | .................... | .................... | .................... |
| Y | 6899 hCoV-19/Romania/USAFSAM-S558/2020\|EPI_ISL_738338\|2 | .................... | ...A................ | ..................... | .................... | .................... | .................... |
| Y | 6900 hCoV-19/Romania/USAFSAM-S559/2020\|EPI_ISL_738339\|2 | .................... | ...A................ | ..................... | .................... | .................... | .................... |
| Y | 6901 hCoV-19/Romania/USAFSAM-S562/2020\|EPI_ISL_738340\|2 | .................... | ...A................ | ..................... | .................... | .................... | .................... |
| Y | 6902 hCoV-19/Romania/USAFSAM-S564/2020\|EPI_ISL_738341\|2 | .................... | .................... | ..................... | .................... | .................... | .................... |
| Y | 6903 hCoV-19/Romania/USAFSAM-S565/2020\|EPI_ISL_738342\|2 | .................... | .................... | ..................... | .................... | .................... | .................... |
| Y | 6904 hCoV-19/Romania/USAFSAM-S569/2020\|EPI_ISL_738343\|2 | .................... | .................... | ..................... | .................... | .................... | .................... |
| Y | 6905 hCoV-19/Romania/USAFSAM-S570/2020\|EPI_ISL_738344\|2 | .................... | .................... | ..................... | .................... | .................... | .................... |

| | | | | | | | | |
|---|---|---|---|---|---|---|---|---|
| Y | 6906 | hCoV-19/Romania/USAFSAM-S571/2020\|EPI_ISL_738345\|2 | ···················· | ····A··············· | ····················· | ···················· | ···················· | ···················· |
| Y | 6907 | hCoV-19/Romania/USAFSAM-S572/2020\|EPI_ISL_738346\|2 | ···················· | ····A··············· | ····················· | ···················· | ···················· | ···················· |
| Y | 6908 | hCoV-19/Romania/USAFSAM-S573/2020\|EPI_ISL_738347\|2 | ···················· | ····A··············· | ····················· | ···················· | ···················· | ···················· |
| Y | 6909 | hCoV-19/Romania/USAFSAM-S575/2020\|EPI_ISL_738348\|2 | ···················· | ···················· | ····················· | ···················· | ···················· | ···················· |
| Y | 6910 | hCoV-19/Romania/USAFSAM-S576/2020\|EPI_ISL_738349\|2 | ···················· | ···················· | ····················· | ···················· | ···················· | ···················· |
| Y | 6911 | hCoV-19/Palestine/AAS27/2020\|EPI_ISL_739660\|2020-1 | ···················· | ···················· | ····················· | ···················· | ···················· | ···················· |
| Y | 6912 | hCoV-19/Palestine/AAS26/2020\|EPI_ISL_739661\|2020-1 | ···················· | ···················· | ····················· | ···················· | ···················· | ···················· |
| Y | 6913 | hCoV-19/Wales/PHWC-49351E/2020\|EPI_ISL_725900\|2020 | ···················· | ···················· | ····················· | ···················· | ···················· | ···················· |
| Y | 6914 | hCoV-19/Wales/PHWC-49352D/2020\|EPI_ISL_725901\|2020 | ···················· | ···················· | ····················· | ···················· | ···················· | ···················· |
| Y | 6915 | hCoV-19/Wales/PHWC-49354B/2020\|EPI_ISL_725903\|2020 | ···················· | ···················· | ····················· | ···················· | ···················· | ···················· |
| Y | 6916 | hCoV-19/Wales/PHWC-49355A/2020\|EPI_ISL_725904\|2020 | ···················· | ···················· | ····················· | ···················· | ···················· | ···················· |
| Y | 6917 | hCoV-19/Wales/PHWC-493578/2020\|EPI_ISL_725906\|2020 | ···················· | ···················· | ····················· | ···················· | ···················· | ···················· |
| Y | 6918 | hCoV-19/Wales/PHWC-493587/2020\|EPI_ISL_725907\|2020 | ···················· | ···················· | ····················· | ···················· | ···················· | ···················· |
| Y | 6919 | hCoV-19/Wales/PHWC-493596/2020\|EPI_ISL_725908\|2020 | ···················· | ···················· | ····················· | ···················· | ···················· | ···················· |
| Y | 6922 | hCoV-19/Wales/PHWC-4935C3/2020\|EPI_ISL_725911\|2020 | ···················· | ···················· | ····················· | ···················· | ···················· | ···················· |
| Y | 6924 | hCoV-19/USA/WI-UW-2334/2020\|EPI_ISL_728100\|2020-11 | ···················· | ···················· | ····················· | ···················· | ···················· | ···················· |
| Y | 6925 | hCoV-19/USA/WI-UW-2335/2020\|EPI_ISL_728101\|2020-11 | ···················· | ·A·················· | ····················· | ···················· | ···················· | ···················· |
| Y | 6926 | hCoV-19/USA/WI-UW-2336/2020\|EPI_ISL_728102\|2020-11 | ···················· | ···················· | ····················· | ···················· | ···················· | ···················· |
| Y | 6929 | hCoV-19/USA/WI-UW-2344/2020\|EPI_ISL_728110\|2020-11 | ···················· | ···················· | ····················· | ···················· | ···················· | ···················· |
| Y | 6930 | hCoV-19/USA/IL-UW-2346/2020\|EPI_ISL_728112\|2020-11 | ···················· | ···················· | ····················· | ···················· | ···················· | ···················· |
| Y | 6931 | hCoV-19/USA/WI-UW-2347/2020\|EPI_ISL_728113\|2020-11 | ···················· | ···················· | ····················· | ···················· | ···················· | ···················· |
| Y | 6933 | hCoV-19/USA/WI-UW-2349/2020\|EPI_ISL_728115\|2020-11 | ···················· | ···················· | ····················· | ···················· | ···················· | ···················· |
| Y | 6934 | hCoV-19/USA/WI-UW-2350/2020\|EPI_ISL_728116\|2020-11 | ···················· | ···················· | ····················· | ···················· | ···················· | ···················· |
| Y | 6937 | hCoV-19/USA/WI-UW-2354/2020\|EPI_ISL_728120\|2020-11 | ···················· | ···················· | ····················· | ···················· | ···················· | ···················· |
| Y | 6940 | hCoV-19/USA/WI-UW-2359/2020\|EPI_ISL_728125\|2020-11 | ···················· | ·A·················· | ····················· | ···················· | ···················· | ···················· |
| Y | 6941 | hCoV-19/USA/WI-UW-2360/2020\|EPI_ISL_728126\|2020-11 | ···················· | ···················· | ····················· | ···················· | ···················· | ···················· |
| Y | 6943 | hCoV-19/USA/WI-UW-2362/2020\|EPI_ISL_728128\|2020-10 | ···················· | ···················· | ····················· | ···················· | ···················· | ···················· |
| Y | 6945 | hCoV-19/USA/WI-UW-2364/2020\|EPI_ISL_728130\|2020-11 | ···················· | ···················· | ····················· | ···················· | ···················· | ···················· |
| Y | 6947 | hCoV-19/USA/WI-UW-2366/2020\|EPI_ISL_728132\|2020-11 | ···················· | ···················· | ····················· | ···················· | ···················· | ···················· |
| Y | 6948 | hCoV-19/USA/WI-UW-2367/2020\|EPI_ISL_728133\|2020-11 | ···················· | ···················· | ····················· | ···················· | ···················· | ···················· |
| Y | 6950 | hCoV-19/USA/WI-UW-2369/2020\|EPI_ISL_728135\|2020-11 | ·····T·············· | ···················· | ····················· | ···················· | ···················· | ···················· |
| Y | 6951 | hCoV-19/USA/WI-UW-2370/2020\|EPI_ISL_728136\|2020-11 | ···················· | ···················· | ····················· | ···················· | ···················· | ···················· |
| Y | 6952 | hCoV-19/USA/WI-UW-2371/2020\|EPI_ISL_728137\|2020-11 | ···················· | ···················· | ····················· | ···················· | ···················· | ···················· |
| Y | 6953 | hCoV-19/USA/WI-UW-2372/2020\|EPI_ISL_728138\|2020-11 | ···················· | ···················· | ····················· | ···················· | ···················· | ···················· |
| Y | 6954 | hCoV-19/USA/WI-UW-2373/2020\|EPI_ISL_728139\|2020-11 | ···················· | ···················· | ····················· | ···················· | ···················· | ···················· |
| Y | 6955 | hCoV-19/USA/WI-UW-2374/2020\|EPI_ISL_728140\|2020-11 | ···················· | ···················· | ····················· | ···················· | ···················· | ···················· |
| Y | 6959 | hCoV-19/USA/WI-UW-2380/2020\|EPI_ISL_728145\|2020-11 | ···················· | ···················· | ····················· | ···················· | ···················· | ···················· |
| Y | 6960 | hCoV-19/USA/WI-UW-2381/2020\|EPI_ISL_728146\|2020-10 | ···················· | ···················· | ····················· | ···················· | ···················· | ···················· |
| Y | 6961 | hCoV-19/USA/WI-UW-2382/2020\|EPI_ISL_728147\|2020-10 | ···················· | ····G··············· | ····················· | ···················· | ···················· | ···················· |
| Y | 6962 | hCoV-19/USA/WI-UW-2383/2020\|EPI_ISL_728148\|2020-10 | ···················· | ···················· | ····················· | ···················· | ···················· | ···················· |
| Y | 6963 | hCoV-19/USA/WI-UW-2384/2020\|EPI_ISL_728149\|2020-10 | ···················· | ···················· | ····················· | ···················· | ···················· | ···················· |
| Y | 6964 | hCoV-19/USA/WI-UW-2387/2020\|EPI_ISL_728152\|2020-10 | ···················· | ···················· | ····················· | ···················· | ···················· | ···················· |
| Y | 6965 | hCoV-19/Japan/FHU-CS56-0301/2020\|EPI_ISL_728153\|20 | ···················· | ···················· | ····················· | ···················· | ···················· | ···················· |
| Y | 6966 | hCoV-19/Japan/FHU-CS77-0222/2020\|EPI_ISL_728154\|20 | ···················· | ···················· | ····················· | ···················· | ···················· | ···················· |
| Y | 6967 | hCoV-19/Japan/FHU-CS84-0222/2020\|EPI_ISL_728155\|20 | ···················· | ···················· | ····················· | ···················· | ···················· | ···················· |
| Y | 6968 | hCoV-19/Japan/FHU-CS108-0222/2020\|EPI_ISL_728156\|2 | ···················· | ···················· | ····················· | ···················· | ···················· | ···················· |
| Y | 6969 | hCoV-19/Japan/FHU-CS108-0224/2020\|EPI_ISL_728159\|2 | ···················· | ···················· | ····················· | ···················· | ···················· | ···················· |
| Y | 6970 | hCoV-19/Malaysia/IMR-WC206980/2020\|EPI_ISL_728165\| | ···················· | ···················· | ····················· | ···················· | ···················· | ···A················ |
| Y | 6971 | hCoV-19/Malaysia/IMR-WC216336/2020\|EPI_ISL_728166\| | ···················· | ·A·················· | ····················· | ···················· | ···················· | ···················· |
| Y | 6972 | hCoV-19/Malaysia/IMR-WC249886/2020\|EPI_ISL_728167\| | ···················· | ·A·················· | ····················· | ···················· | ···················· | ···A················ |
| Y | 6973 | hCoV-19/Malaysia/IMR-WC253038/2020\|EPI_ISL_728169\| | ···················· | ·A·················· | ····················· | ···················· | ···················· | ···················· |
| Y | 6974 | hCoV-19/Malaysia/IMR-WC253176/2020\|EPI_ISL_728170\| | ···················· | ·A·················· | ····················· | ···················· | ···················· | ···A················ |
| Y | 6975 | hCoV-19/Malaysia/IMR-WC257003/2020\|EPI_ISL_728178\| | ···················· | ·A·················· | ····················· | ···················· | ···················· | ···················· |
| Y | 6976 | hCoV-19/Singapore/1443/2020\|EPI_ISL_728179\|2020-12 | ···················· | ·A·················· | ····················· | ···················· | ···················· | ···················· |
| Y | 6977 | hCoV-19/Singapore/1444/2020\|EPI_ISL_728180\|2020-12 | ···················· | ·A·················· | ····················· | ···················· | ···················· | ···A················ |
| Y | 6978 | hCoV-19/Singapore/1445/2020\|EPI_ISL_728181\|2020-12 | ···················· | ···················· | ····················· | ···················· | ···················· | ···A················ |
| Y | 6979 | hCoV-19/Singapore/1446/2020\|EPI_ISL_728182\|2020-12 | ···················· | ·A·················· | ····················· | ···················· | ···················· | ···················· |
| Y | 6980 | hCoV-19/Singapore/1447/2020\|EPI_ISL_728183\|2020-12 | ···················· | ···················· | ····················· | ···················· | ···················· | ···················· |
| Y | 6981 | hCoV-19/Singapore/1448/2020\|EPI_ISL_728184\|2020-12 | ···················· | ···················· | ····················· | ···················· | ···················· | ···················· |
| Y | 6982 | hCoV-19/Singapore/1449/2020\|EPI_ISL_728185\|2020-12 | ···················· | ·A·················· | ····················· | ···················· | ···················· | ···················· |
| Y | 6983 | hCoV-19/Singapore/1450/2020\|EPI_ISL_728186\|2020-12 | ···················· | ···················· | ·······R············· | ···················· | ···················· | ···A················ |
| Y | 6984 | hCoV-19/Singapore/1451/2020\|EPI_ISL_728187\|2020-12 | ···················· | ···················· | ····················· | ···················· | ···················· | ···················· |
| Y | 6985 | hCoV-19/Singapore/1452/2020\|EPI_ISL_728188\|2020-12 | ···················· | ···················· | ····················· | ···················· | ···················· | ···················· |
| Y | 6986 | hCoV-19/Germany/ChVir8135/2020\|EPI_ISL_729415\|2020 | ···················· | ···················· | ····················· | ···················· | ···················· | ···················· |
| Y | 6987 | hCoV-19/Germany/DE-SH-ChVir20806/2020\|EPI_ISL_7294 | ···················· | ···················· | ····················· | ···················· | ···················· | ···················· |
| Y | 6988 | hCoV-19/Germany/DE-SH-ChVir20054/2020\|EPI_ISL_7294 | ···················· | ···················· | ····················· | ···················· | ···················· | ···················· |
| Y | 6990 | hCoV-19/Germany/DE-SH-ChVir9283/2020\|EPI_ISL_72941 | ···················· | ·A·················· | ····················· | ···················· | ···················· | ···A················ |
| Y | 6991 | hCoV-19/Germany/DE-SH-ChVir20770/2020\|EPI_ISL_7294 | ···················· | ···················· | ····················· | ···················· | ···················· | ···················· |
| Y | 6992 | hCoV-19/Germany/DE-SH-ChVir9290/2020\|EPI_ISL_72942 | ···················· | ···················· | ····················· | ···················· | ···················· | ···················· |
| Y | 6993 | hCoV-19/Germany/DE-SH-ChVir9632/2020\|EPI_ISL_72942 | ···················· | ···················· | ····················· | ···················· | ···················· | ···················· |
| Y | 6994 | hCoV-19/Germany/DE-SH-ChVir9633/2020\|EPI_ISL_72942 | ···················· | ···················· | ····················· | ···················· | ···················· | ···················· |
| Y | 6995 | hCoV-19/Germany/DE-SH-ChVir9933/2020\|EPI_ISL_72942 | ···················· | ···················· | ····················· | ···················· | ···················· | ···················· |
| Y | 6996 | hCoV-19/Germany/DE-SH-ChVir9934/2020\|EPI_ISL_72942 | ···················· | ···················· | ····················· | ···················· | ···················· | ···················· |
| Y | 6997 | hCoV-19/Germany/DE-SH-ChVir9939/2020\|EPI_ISL_72942 | ···················· | ···················· | ····················· | ···················· | ···················· | ···················· |
| Y | 6998 | hCoV-19/Germany/DE-SH-ChVir9944/2020\|EPI_ISL_72942 | ···················· | ···················· | ····················· | ···················· | ···················· | ···················· |
| Y | 6999 | hCoV-19/Germany/DE-SH-ChVir9951/2020\|EPI_ISL_72942 | ···················· | ···················· | ····················· | ···················· | ···················· | ···················· |
| Y | 7000 | hCoV-19/Germany/DE-SH-ChVir9953/2020\|EPI_ISL_72942 | ···················· | ···················· | ····················· | ···················· | ···················· | ···················· |
| Y | 7001 | hCoV-19/Germany/DE-SH-ChVir20041/2020\|EPI_ISL_7294 | ···················· | ···················· | ····················· | ···················· | ···················· | ···················· |
| Y | 7002 | hCoV-19/Germany/DE-SH-ChVir20298/2020\|EPI_ISL_7294 | ···················· | ···················· | ····················· | ···················· | ···················· | ···················· |
| Y | 7003 | hCoV-19/Germany/DE-SH-ChVir20303/2020\|EPI_ISL_7294 | ···················· | ···················· | ····················· | ···················· | ···················· | ···················· |

| | | | | | | | |
|---|---|---|---|---|---|---|---|
| Y | 7004 hCoV-19/Germany/DE-SH-ChVir20309/2020\|EPI_ISL_7294 | ···················· | ···················· | ···················· | ···················· | ···················· | ···················· |
| Y | 7005 hCoV-19/Germany/DE-SH-ChVir9981/2020\|EPI_ISL_72944 | ···················· | ···················· | ···················· | ···················· | ···················· | ···················· |
| Y | 7006 hCoV-19/Germany/DE-SH-ChVir20747/2020\|EPI_ISL_7294 | ···················· | ···················· | ···················· | ···················· | ···················· | ···················· |
| Y | 7007 hCoV-19/Germany/DE-SH-ChVir20749/2020\|EPI_ISL_7294 | ···················· | ···················· | ···················· | ···················· | ···················· | ···················· |
| Y | 7008 hCoV-19/Germany/DE-SH-ChVir20772/2020\|EPI_ISL_7294 | ···················· | ···················· | ···················· | ···················· | ···················· | ···················· |
| Y | 7009 hCoV-19/Germany/DE-SH-ChVir20774/2020\|EPI_ISL_7294 | ···················· | ···················· | ···················· | ···················· | ···················· | ···················· |
| Y | 7010 hCoV-19/Germany/DE-SH-ChVir20791/2020\|EPI_ISL_7294 | ···················· | ···················· | ···················· | ···················· | ···················· | ···················· |
| Y | 7011 hCoV-19/Germany/DE-SH-ChVir20802/2020\|EPI_ISL_7294 | ···················· | ···················· | ···················· | ···················· | ···················· | ···················· |
| Y | 7012 hCoV-19/Germany/DE-SH-ChVir20805/2020\|EPI_ISL_7294 | ···················· | ···················· | ···················· | ···················· | ···················· | ···················· |
| Y | 7013 hCoV-19/Germany/DE-SH-ChVir20831/2020\|EPI_ISL_7294 | ···················· | ···················· | ···················· | ···················· | ···················· | ···················· |
| Y | 7014 hCoV-19/Germany/DE-SH-ChVir20813/2020\|EPI_ISL_7294 | ···················· | ···················· | ···················· | ···················· | ···················· | ···················· |
| Y | 7015 hCoV-19/Germany/DE-SH-ChVir20814/2020\|EPI_ISL_7294 | ···················· | ···················· | ···················· | ···················· | ···················· | ···················· |
| Y | 7016 hCoV-19/Germany/DE-SH-ChVir20817/2020\|EPI_ISL_7294 | ···················· | ···················· | ···················· | ···················· | ···················· | ···················· |
| Y | 7017 hCoV-19/Germany/DE-SH-ChVir20821/2020\|EPI_ISL_7294 | ···················· | ···················· | ···················· | ···················· | ···················· | ···················· |
| Y | 7018 hCoV-19/Germany/DE-SH-ChVir20843/2020\|EPI_ISL_7294 | ···················· | ···················· | ···················· | ···················· | ···················· | ···················· |
| Y | 7019 hCoV-19/Germany/DE-SH-ChVir20844/2020\|EPI_ISL_7294 | ···················· | ···················· | ···················· | ···················· | ···················· | ···················· |
| Y | 7020 hCoV-19/Germany/DE-SH-ChVir20834/2020\|EPI_ISL_7294 | ···················· | ···················· | ···················· | ···················· | ···················· | ···················· |
| Y | 7021 hCoV-19/Germany/DE-SH-ChVir3199/2020\|EPI_ISL_72946 | ···················· | ···················· | ···················· | ···················· | ···················· | ···················· |
| Y | 7022 hCoV-19/Germany/DE-SH-ChVir9943/2020\|EPI_ISL_72946 | ···················· | ···················· | ···················· | ···················· | ···················· | ···················· |
| Y | 7023 hCoV-19/Germany/DE-SH-ChVir20810/2020\|EPI_ISL_7294 | ···················· | ···················· | ···················· | ···················· | ···················· | ···················· |
| Y | 7024 hCoV-19/Germany/DE-SH-ChVir3210/2020\|EPI_ISL_72946 | ···················· | ···················· | ···················· | ···················· | ···················· | ···················· |
| Y | 7025 hCoV-19/Germany/DE-BW-ChVir1600/2020\|EPI_ISL_72947 | ···················· | ···················· | ···················· | ···················· | ···················· | ···················· |
| Y | 7026 hCoV-19/Germany/DE-RP-ChVir1672/2020\|EPI_ISL_72947 | ···················· | ···················· | ···················· | ···················· | ···················· | ···················· |
| Y | 7027 hCoV-19/Germany/DE-BW-ChVir1611/2020\|EPI_ISL_72947 | ···················· | ···················· | ···················· | ···················· | ···················· | ···················· |
| Y | 7028 hCoV-19/Germany/DE-NW-ChVir1671/2020\|EPI_ISL_72947 | ···················· | ···················· | ···················· | ···················· | ···················· | ···················· |
| Y | 7029 hCoV-19/Germany/DE-BE-ChVir1670/2020\|EPI_ISL_72947 | ···················· | ···················· | ···················· | ···················· | ···················· | ···················· |
| Y | 7030 hCoV-19/Germany/DE-NW-ChVir1683/2020\|EPI_ISL_72947 | ···················· | ···················· | ···················· | ···················· | ···················· | ···················· |
| Y | 7031 hCoV-19/Germany/DE-BE-ChVir1685/2020\|EPI_ISL_72947 | ···················· | ···················· | ···················· | ···················· | ···················· | ···················· |
| Y | 7032 hCoV-19/Germany/DE-MV-ChVir1689/2020\|EPI_ISL_72948 | ···················· | ···················· | ···················· | ···················· | ···················· | ···················· |
| Y | 7033 hCoV-19/Germany/DE-BE-ChVir1716/2020\|EPI_ISL_72948 | ···················· | ···················· | ···················· | ···················· | ···················· | ···················· |
| Y | 7034 hCoV-19/Germany/DE-BE-ChVir1897/2020\|EPI_ISL_72948 | ···················· | ···················· | ···················· | ···················· | ···················· | ···················· |
| Y | 7035 hCoV-19/Germany/DE-BE-ChVir1962/2020\|EPI_ISL_72949 | ···················· | ···················· | ···················· | ···················· | ···················· | ···················· |
| Y | 7036 hCoV-19/Germany/ChVir1967/2020\|EPI_ISL_729491\|2020 | ···················· | ···················· | ···················· | ···················· | ···················· | ···················· |
| Y | 7037 hCoV-19/Germany/DE-BE-ChVir1968/2020\|EPI_ISL_72949 | ···················· | ···················· | ···················· | ···················· | ···················· | ···················· |
| Y | 7038 hCoV-19/Germany/DE-BE-ChVir1970/2020\|EPI_ISL_72949 | ···················· | ···················· | ···················· | ···················· | ···················· | ···················· |
| Y | 7039 hCoV-19/Germany/DE-BY-ChVir1980/2020\|EPI_ISL_72949 | ···················· | ···················· | ···················· | ···················· | ···················· | ···················· |
| Y | 7040 hCoV-19/Germany/DE-BY-ChVir1981/2020\|EPI_ISL_72949 | ···················· | ···················· | ···················· | ···················· | ···················· | ···················· |
| Y | 7041 hCoV-19/Germany/ChVir1993/2020\|EPI_ISL_729496\|2020 | ···················· | ···················· | ···················· | ···················· | ···················· | ···················· |
| Y | 7042 hCoV-19/Germany/ChVir1994/2020\|EPI_ISL_729497\|2020 | ···················· | ···················· | ···················· | ···················· | ···················· | ···················· |
| Y | 7043 hCoV-19/Germany/DE-SH-ChVir2000/2020\|EPI_ISL_72949 | ···················· | ···················· | ···················· | ···················· | ···················· | ···················· |
| Y | 7044 hCoV-19/Germany/DE-BE-ChVir2192/2020\|EPI_ISL_72949 | ···················· | ···················· | ···················· | ···················· | ···················· | ···················· |
| Y | 7045 hCoV-19/Jordan/AM-ALSR-5162/2020\|EPI_ISL_730401\|20 | ···················· | ···················· | ···················· | ···················· | ···················· | ···················· |
| Y | 7046 hCoV-19/Jordan/AM-ALSR-5163/2020\|EPI_ISL_730402\|20 | ···················· | ···················· | ···················· | ···················· | ···················· | ···················· |
| Y | 7047 hCoV-19/Jordan/AM-ALSR-5164/2020\|EPI_ISL_730403\|20 | ···················· | ···················· | ···················· | ···················· | ···················· | ···················· |
| Y | 7048 hCoV-19/Jordan/AM-ALSR-5167/2020\|EPI_ISL_730406\|20 | ···················· | ···················· | ···················· | ···················· | ···················· | ···················· |
| Y | 7049 hCoV-19/Jordan/AM-ALSR-5168/2020\|EPI_ISL_730407\|20 | ···················· | ···················· | ···················· | ···················· | ······T············· | ···················· |
| Y | 7050 hCoV-19/Jordan/AM-ALSR-5169/2020\|EPI_ISL_730408\|20 | ···················· | ···················· | ···················· | ···················· | ···················· | ···················· |
| Y | 7051 hCoV-19/Jordan/AM-ALSR-5170/2020\|EPI_ISL_730409\|20 | ···················· | ···················· | ···················· | ···················· | ···················· | ···················· |
| Y | 7052 hCoV-19/Jordan/AM-ALSR-5171/2020\|EPI_ISL_730410\|20 | ···················· | ···················· | ···················· | ···················· | ···················· | ···················· |
| Y | 7053 hCoV-19/Jordan/AM-ALSR-5172/2020\|EPI_ISL_730411\|20 | ···················· | ···················· | ···················· | ···················· | ···················· | ···················· |
| Y | 7054 hCoV-19/Jordan/AM-ALSR-5173/2020\|EPI_ISL_730412\|20 | ···················· | ·A·················· | ···················· | ···················· | ···················· | ··A················· |
| Y | 7055 hCoV-19/Jordan/AM-ALSR-5185/2020\|EPI_ISL_730424\|20 | ···················· | ···················· | ···················· | ···················· | ···················· | ···················· |
| Y | 7056 hCoV-19/Jordan/AM-ALSR-5186/2020\|EPI_ISL_730425\|20 | ···················· | ···················· | ···················· | ···················· | ···················· | ···················· |
| Y | 7057 hCoV-19/Jordan/AM-ALSR-5187/2020\|EPI_ISL_730426\|20 | ···················· | ·A·················· | ···················· | ···················· | ···················· | ··A················· |
| Y | 7058 hCoV-19/Jordan/AM-ALSR-5188/2020\|EPI_ISL_730427\|20 | ···················· | ·A·················· | ···················· | ···················· | ···················· | ··A················· |
| Y | 7059 hCoV-19/Jordan/AM-ALSR-5189/2020\|EPI_ISL_730428\|20 | ···················· | ···················· | ···················· | ···················· | ···················· | ···················· |
| Y | 7060 hCoV-19/Jordan/IR-ALSR-5190/2020\|EPI_ISL_730429\|20 | ···················· | ···················· | ···················· | ···················· | ···················· | ···················· |
| Y | 7061 hCoV-19/Jordan/IR-ALSR-5192/2020\|EPI_ISL_730431\|20 | ···················· | ·A·················· | ···················· | ···················· | ···················· | ··A················· |
| Y | 7062 hCoV-19/Jordan/IR-ALSR-5194/2020\|EPI_ISL_730432\|20 | ···················· | ···················· | ···················· | ···················· | ···················· | ···················· |
| Y | 7063 hCoV-19/Jordan/IR-ALSR-5195/2020\|EPI_ISL_730433\|20 | ···················· | ···················· | ···················· | ···················· | ···················· | ···················· |
| Y | 7064 hCoV-19/Jordan/IR-ALSR-5196/2020\|EPI_ISL_730434\|20 | ···················· | ·A·················· | ···················· | ···················· | ···················· | ··A················· |
| Y | 7065 hCoV-19/Jordan/IR-ALSR-5197/2020\|EPI_ISL_730435\|20 | ···················· | ···················· | ···················· | ···················· | ···················· | ···················· |
| Y | 7066 hCoV-19/Jordan/IR-ALSR-5198/2020\|EPI_ISL_730436\|20 | ···················· | ···················· | ···················· | ···················· | ···················· | ···················· |
| Y | 7067 hCoV-19/Jordan/IR-ALSR-5199/2020\|EPI_ISL_730437\|20 | ···················· | ···················· | ···················· | ···················· | ···················· | ···················· |
| Y | 7068 hCoV-19/Jordan/IR-ALSR-5200/2020\|EPI_ISL_730438\|20 | ···················· | ···················· | ···················· | ···················· | ···················· | ···················· |
| Y | 7069 hCoV-19/Jordan/IR-ALSR-5201/2020\|EPI_ISL_730439\|20 | ···················· | ···················· | ···················· | ···················· | ···················· | ···················· |
| Y | 7070 hCoV-19/Jordan/IR-ALSR-5202/2020\|EPI_ISL_730440\|20 | ···················· | ···················· | ···················· | ···················· | ···················· | ···················· |
| Y | 7071 hCoV-19/Jordan/IR-ALSR-5203/2020\|EPI_ISL_730441\|20 | ···················· | ···················· | ···················· | ···················· | ···················· | ···················· |
| Y | 7072 hCoV-19/Jordan/IR-ALSR-5204/2020\|EPI_ISL_730442\|20 | ···················· | ···················· | ···················· | ···················· | ···················· | ···················· |
| Y | 7073 hCoV-19/Jordan/IR-ALSR-5205/2020\|EPI_ISL_730443\|20 | ···················· | ···················· | ···················· | ···················· | ···················· | ···················· |
| Y | 7074 hCoV-19/Jordan/IR-ALSR-5206/2020\|EPI_ISL_730444\|20 | ···················· | ···················· | ···················· | ···················· | ···················· | ···················· |
| Y | 7075 hCoV-19/Jordan/IR-ALSR-5207/2020\|EPI_ISL_730445\|20 | ···················· | ·A·················· | ···················· | ···················· | ···················· | ··A················· |
| Y | 7076 hCoV-19/Jordan/IR-ALSR-5208/2020\|EPI_ISL_730446\|20 | ···················· | ···················· | ···················· | ···················· | ···················· | ···················· |
| Y | 7077 hCoV-19/Jordan/IR-ALSR-5209/2020\|EPI_ISL_730447\|20 | ···················· | ···················· | ···················· | ···················· | ···················· | ···················· |
| Y | 7078 hCoV-19/Jordan/IR-ALSR-5210/2020\|EPI_ISL_730448\|20 | ···················· | ···················· | ···················· | ···················· | ···················· | ···················· |
| Y | 7079 hCoV-19/Jordan/IR-ALSR-5211/2020\|EPI_ISL_730449\|20 | ···················· | ···················· | ···················· | ···················· | ···················· | ···················· |
| Y | 7080 hCoV-19/Jordan/IR-ALSR-5212/2020\|EPI_ISL_730450\|20 | ···················· | ···················· | ···················· | ···················· | ···················· | ···················· |
| Y | 7081 hCoV-19/Jordan/IR-ALSR-5213/2020\|EPI_ISL_730451\|20 | ···················· | ·A·················· | ···················· | ···················· | ···················· | ··A················· |
| Y | 7082 hCoV-19/Jordan/IR-ALSR-5214/2020\|EPI_ISL_730452\|20 | ···················· | ···················· | ···················· | ···················· | ···················· | ···················· |
| Y | 7083 hCoV-19/Jordan/IR-ALSR-5215/2020\|EPI_ISL_730453\|20 | ···················· | ···················· | ···················· | ···················· | ···················· | ···················· |

| | | | | | | | | |
|---|---|---|---|---|---|---|---|---|
| Y | 7084 | hCoV-19/Jordan/IR-ALSR-5216/2020\|EPI_ISL_730454\|20 | ···················· | ·A·················· | ···················· | ···················· | ·········T·········· | ···A················ |
| Y | 7085 | hCoV-19/Jordan/IR-ALSR-5217/2020\|EPI_ISL_730455\|20 | ···················· | ···················· | ···················· | ···················· | ···················· | ···················· |
| Y | 7086 | hCoV-19/Jordan/IR-ALSR-5218/2020\|EPI_ISL_730456\|20 | ···················· | ···················· | ···················· | ···················· | ···················· | ···················· |
| Y | 7087 | hCoV-19/Jordan/IR-ALSR-5219/2020\|EPI_ISL_730457\|20 | ···················· | ···················· | ···················· | ···················· | ···················· | ···················· |
| Y | 7088 | hCoV-19/Jordan/IR-ALSR-5220/2020\|EPI_ISL_730458\|20 | ···················· | ···················· | ···················· | ···················· | ···················· | ···················· |
| Y | 7089 | hCoV-19/Jordan/IR-ALSR-5221/2020\|EPI_ISL_730459\|20 | ···················· | ···················· | ···················· | ···················· | ···················· | ···················· |
| Y | 7090 | hCoV-19/Jordan/IR-ALSR-5223/2020\|EPI_ISL_730461\|20 | ···················· | ···················· | ···················· | ···················· | ···················· | ···················· |
| Y | 7091 | hCoV-19/Jordan/IR-ALSR-5224/2020\|EPI_ISL_730462\|20 | ···················· | ···················· | ···················· | ···················· | ···················· | ···················· |
| Y | 7092 | hCoV-19/Jordan/IR-ALSR-5225/2020\|EPI_ISL_730463\|20 | ···················· | ···················· | ···················· | ···················· | ···················· | ···················· |
| Y | 7093 | hCoV-19/Jordan/IR-ALSR-5226/2020\|EPI_ISL_730464\|20 | ···················· | ···················· | ···················· | ···················· | ···················· | ···················· |
| Y | 7094 | hCoV-19/Jordan/AZ-ALSR-5227/2020\|EPI_ISL_730465\|20 | ···················· | ···················· | ···················· | ···················· | ···················· | ···················· |
| Y | 7095 | hCoV-19/Jordan/IR-ALSR-5228/2020\|EPI_ISL_730466\|20 | ···················· | ···················· | ···················· | ···················· | ···················· | ···················· |
| Y | 7096 | hCoV-19/Jordan/IR-ALSR-5229/2020\|EPI_ISL_730467\|20 | ···················· | ·A·················· | ···················· | ···················· | ···················· | ···A················ |
| Y | 7097 | hCoV-19/Jordan/IR-ALSR-5230/2020\|EPI_ISL_730468\|20 | ···················· | ···················· | ···················· | ···················· | ···················· | ···················· |
| Y | 7098 | hCoV-19/Jordan/IR-ALSR-5231/2020\|EPI_ISL_730469\|20 | ···················· | ···················· | ···················· | ···················· | ···················· | ···················· |
| Y | 7099 | hCoV-19/Jordan/AQ-ALSR-5241/2020\|EPI_ISL_730479\|20 | ···················· | ···················· | ···················· | ···················· | ···················· | ···················· |
| Y | 7100 | hCoV-19/Jordan/AQ-ALSR-5242/2020\|EPI_ISL_730480\|20 | ···················· | ···················· | ···················· | ···················· | ···················· | ···················· |
| Y | 7101 | hCoV-19/Jordan/AQ-ALSR-5243/2020\|EPI_ISL_730481\|20 | ···················· | ···················· | ···················· | ···················· | ···················· | ···················· |
| Y | 7102 | hCoV-19/Jordan/AQ-ALSR-5244/2020\|EPI_ISL_730482\|20 | ···················· | ···················· | ···················· | ···················· | ···················· | ···················· |
| Y | 7103 | hCoV-19/Jordan/AQ-ALSR-5245/2020\|EPI_ISL_730483\|20 | ···················· | ···················· | ···················· | ···················· | ···················· | ···················· |
| Y | 7104 | hCoV-19/Jordan/AQ-ALSR-5246/2020\|EPI_ISL_730484\|20 | ···················· | ···················· | ···················· | ···················· | ···················· | ···················· |
| Y | 7105 | hCoV-19/Jordan/AQ-ALSR-5247/2020\|EPI_ISL_730485\|20 | ···················· | ···················· | ···················· | ···················· | ···················· | ···················· |
| Y | 7106 | hCoV-19/Jordan/AQ-ALSR-5248/2020\|EPI_ISL_730486\|20 | ···················· | ···················· | ···················· | ···················· | ···················· | ···················· |
| Y | 7107 | hCoV-19/Jordan/AQ-ALSR-5249/2020\|EPI_ISL_730487\|20 | ···················· | ···················· | ···················· | ···················· | ···················· | ···················· |
| Y | 7108 | hCoV-19/Jordan/AQ-ALSR-5250/2020\|EPI_ISL_730488\|20 | ···················· | ···················· | ···················· | ···················· | ···················· | ···················· |
| Y | 7109 | hCoV-19/Jordan/AQ-ALSR-5251/2020\|EPI_ISL_730489\|20 | ···················· | ···················· | ···················· | ···················· | ···················· | ···················· |
| Y | 7110 | hCoV-19/Jordan/AQ-ALSR-5252/2020\|EPI_ISL_730490\|20 | ···················· | ···················· | ···················· | ···················· | ···················· | ···················· |
| Y | 7111 | hCoV-19/Jordan/AQ-ALSR-5253/2020\|EPI_ISL_730491\|20 | ···················· | ···················· | ···················· | ···················· | ···················· | ···················· |
| Y | 7112 | hCoV-19/Jordan/AQ-ALSR-5254/2020\|EPI_ISL_730492\|20 | ···················· | ···················· | ···················· | ···················· | ···················· | ···················· |
| Y | 7113 | hCoV-19/Jordan/AQ-ALSR-5255/2020\|EPI_ISL_730493\|20 | ···················· | ···················· | ···················· | ···················· | ···················· | ···················· |
| Y | 7114 | hCoV-19/Jordan/AQ-ALSR-5256/2020\|EPI_ISL_730494\|20 | ···················· | ···················· | ···················· | ···················· | ···················· | ···················· |
| Y | 7115 | hCoV-19/Jordan/AQ-ALSR-5257/2020\|EPI_ISL_730495\|20 | ···················· | ···················· | ···················· | ···················· | ···················· | ···················· |
| Y | 7116 | hCoV-19/Jordan/AQ-ALSR-5258/2020\|EPI_ISL_730496\|20 | ···················· | ···················· | ···················· | ···················· | ···················· | ···················· |
| Y | 7117 | hCoV-19/Jordan/AQ-ALSR-5259/2020\|EPI_ISL_730497\|20 | ···················· | ···················· | ···················· | ···················· | ···················· | ···················· |
| Y | 7118 | hCoV-19/Jordan/AQ-ALSR-5260/2020\|EPI_ISL_730498\|20 | ···················· | ···················· | ···················· | ···················· | ···················· | ···················· |
| Y | 7119 | hCoV-19/Jordan/AQ-ALSR-5261/2020\|EPI_ISL_730499\|20 | ···················· | ···················· | ···················· | ···················· | ···················· | ···················· |
| Y | 7120 | hCoV-19/England/ALDP-C3AA21/2020\|EPI_ISL_731711\|20 | ···················· | ···················· | ···················· | ···················· | ···················· | ···················· |
| Y | 7121 | hCoV-19/England/ALDP-C3CBD1/2020\|EPI_ISL_731712\|20 | ···················· | ···················· | ···················· | ···················· | ···················· | ···················· |
| Y | 7122 | hCoV-19/England/ALDP-C3CD08/2020\|EPI_ISL_731713\|20 | ···················· | ···················· | ···················· | ···················· | ···················· | ···················· |
| Y | 7123 | hCoV-19/England/ALDP-C3B325/2020\|EPI_ISL_731714\|20 | ···················· | ···················· | ···················· | ···················· | ···················· | ···················· |
| Y | 7124 | hCoV-19/England/ALDP-C3C6C1/2020\|EPI_ISL_731717\|20 | ···················· | ···················· | ···················· | ···················· | ···················· | ···················· |
| Y | 7125 | hCoV-19/England/ALDP-C3D10B/2020\|EPI_ISL_731718\|20 | ···················· | ···················· | ···················· | ···················· | ···················· | ···················· |
| Y | 7126 | hCoV-19/England/ALDP-C3B4B9/2020\|EPI_ISL_731719\|20 | ···················· | ···················· | ···················· | ···················· | ···················· | ···················· |
| Y | 7127 | hCoV-19/England/ALDP-C3D54B/2020\|EPI_ISL_731720\|20 | ···················· | ···················· | ···················· | ···················· | ···················· | ···················· |
| Y | 7128 | hCoV-19/England/ALDP-C3ABF1/2020\|EPI_ISL_731724\|20 | ···················· | ···················· | ···················· | ···················· | ···················· | ···················· |
| Y | 7129 | hCoV-19/England/ALDP-C3BC75/2020\|EPI_ISL_731726\|20 | ···················· | ···················· | ···················· | ···················· | ···················· | ···················· |
| Y | 7130 | hCoV-19/England/ALDP-C3C078/2020\|EPI_ISL_731727\|20 | ···················· | ···················· | ···················· | ···················· | ···················· | ···················· |
| Y | 7131 | hCoV-19/England/ALDP-C3BD36/2020\|EPI_ISL_731728\|20 | ···················· | ···················· | ···················· | ···················· | ···················· | ···················· |
| Y | 7132 | hCoV-19/England/ALDP-C3B079/2020\|EPI_ISL_731731\|20 | ···················· | ···················· | ···················· | ···················· | ···················· | ···················· |
| Y | 7133 | hCoV-19/England/ALDP-C3CBC2/2020\|EPI_ISL_731732\|20 | ···················· | ···················· | ···················· | ···················· | ···················· | ···················· |
| Y | 7134 | hCoV-19/England/ALDP-C3D6EE/2020\|EPI_ISL_731733\|20 | ···················· | ···················· | ···················· | ···················· | ···················· | ···················· |
| Y | 7135 | hCoV-19/England/ALDP-C3D569/2020\|EPI_ISL_731734\|20 | ···················· | ···················· | ···················· | ···················· | ···················· | ···················· |
| Y | 7136 | hCoV-19/England/ALDP-C3CE9C/2020\|EPI_ISL_731738\|20 | ···················· | ···················· | ···················· | ···················· | ···················· | ···················· |
| Y | 7137 | hCoV-19/England/ALDP-C3BE15/2020\|EPI_ISL_731740\|20 | ···················· | ···················· | ···················· | ···················· | ···················· | ···················· |
| Y | 7138 | hCoV-19/England/ALDP-C3D174/2020\|EPI_ISL_731741\|20 | ···················· | ···················· | ···················· | ···················· | ···················· | ···················· |
| Y | 7139 | hCoV-19/England/ALDP-C3CFF3/2020\|EPI_ISL_731742\|20 | ···················· | ···················· | ···················· | ···················· | ···················· | ···················· |
| Y | 7140 | hCoV-19/England/ALDP-C3D727/2020\|EPI_ISL_731743\|20 | ···················· | ···················· | ···················· | ···················· | ···················· | ···················· |
| Y | 7141 | hCoV-19/England/ALDP-C3CC0B/2020\|EPI_ISL_731755\|20 | ···················· | ···················· | ···················· | ···················· | ···················· | ···················· |
| Y | 7142 | hCoV-19/England/ALDP-C3BC93/2020\|EPI_ISL_731756\|20 | ···················· | ···················· | ···················· | ···················· | ···················· | ···················· |
| Y | 7143 | hCoV-19/England/ALDP-C3C0B4/2020\|EPI_ISL_731758\|20 | ···················· | ···················· | ···················· | ···················· | ···················· | ···················· |
| Y | 7144 | hCoV-19/England/ALDP-C3BB4B/2020\|EPI_ISL_731759\|20 | ···················· | ···················· | ···················· | ···················· | ···················· | ···················· |
| Y | 7145 | hCoV-19/England/ALDP-C3CB4A/2020\|EPI_ISL_731760\|20 | ···················· | ···················· | ···················· | ···················· | ···················· | ···················· |
| Y | 7146 | hCoV-19/England/ALDP-C3D675/2020\|EPI_ISL_731761\|20 | ···················· | ···················· | ···················· | ···················· | ···················· | ···················· |
| Y | 7147 | hCoV-19/England/ALDP-C3CFC6/2020\|EPI_ISL_731762\|20 | ···················· | ···················· | ···················· | ···················· | ···A················ | ···················· |
| Y | 7148 | hCoV-19/England/ALDP-C3B167/2020\|EPI_ISL_731763\|20 | ···················· | ···················· | ···················· | ···················· | ···················· | ···················· |
| Y | 7149 | hCoV-19/England/ALDP-C3D192/2020\|EPI_ISL_731765\|20 | ···················· | ···················· | ···················· | ···················· | ···················· | ···················· |
| Y | 7150 | hCoV-19/England/ALDP-C3A8EB/2020\|EPI_ISL_731767\|20 | ···················· | ···················· | ···················· | ···················· | ······T············· | ···················· |
| Y | 7151 | hCoV-19/England/ALDP-C3BB3C/2020\|EPI_ISL_731768\|20 | ···················· | ···················· | ···················· | ···················· | ···················· | ···················· |
| Y | 7152 | hCoV-19/England/ALDP-C3B0D3/2020\|EPI_ISL_731770\|20 | ···················· | ···················· | ···················· | ···················· | ···················· | ···················· |
| Y | 7153 | hCoV-19/England/ALDP-C3D6C0/2020\|EPI_ISL_731771\|20 | ···················· | ···················· | ···················· | ···················· | ···················· | ···················· |
| Y | 7154 | hCoV-19/England/ALDP-C3BC48/2020\|EPI_ISL_731772\|20 | ···················· | ···················· | ···················· | ···················· | ···················· | ···················· |
| Y | 7155 | hCoV-19/England/ALDP-C3BF4F/2020\|EPI_ISL_731774\|20 | ···················· | ···················· | ···················· | ···················· | ···················· | ···················· |
| Y | 7156 | hCoV-19/England/ALDP-C3D11A/2020\|EPI_ISL_731777\|20 | ···················· | ···················· | ···················· | ···················· | ···················· | ···················· |
| Y | 7157 | hCoV-19/England/ALDP-C3D147/2020\|EPI_ISL_731779\|20 | ···················· | ···················· | ···················· | ···················· | ···················· | ···················· |
| Y | 7158 | hCoV-19/England/ALDP-C3D29F/2020\|EPI_ISL_731780\|20 | ···················· | ···················· | ···················· | ···················· | ···················· | ···················· |
| Y | 7159 | hCoV-19/England/ALDP-C3DE22/2020\|EPI_ISL_731781\|20 | ···················· | ·A·················· | ···················· | ···················· | ···················· | ···A················ |
| Y | 7160 | hCoV-19/England/ALDP-C3BE33/2020\|EPI_ISL_731782\|20 | ···················· | ···················· | ···················· | ···················· | ···················· | ···················· |
| Y | 7161 | hCoV-19/England/ALDP-C3BDDC/2020\|EPI_ISL_731784\|20 | ···················· | ···················· | ···················· | ···················· | ···················· | ···················· |
| Y | 7162 | hCoV-19/England/ALDP-C3B862/2020\|EPI_ISL_731786\|20 | ···················· | ···················· | ···················· | ···················· | ···················· | ···················· |
| Y | 7163 | hCoV-19/England/ALDP-C3B246/2020\|EPI_ISL_731788\|20 | ···················· | ···················· | ···················· | ···················· | ···················· | ···················· |

| | | | | | | | | | |
|---|---|---|---|---|---|---|---|---|---|
| Y | 7164 | hCoV-19/England/ALDP-C3BCB1/2020\|EPI_ISL_731789\|20 | .................... | .................... | .................... | .................... | .................... | .................... | |
| Y | 7165 | hCoV-19/England/ALDP-C3DD16/2020\|EPI_ISL_731793\|20 | .................... | .................... | .................... | .................... | .................... | .................... | |
| Y | 7166 | hCoV-19/England/ALDP-C3D781/2020\|EPI_ISL_731794\|20 | .................... | .................... | .................... | .................... | .................... | .................... | |
| Y | 7167 | hCoV-19/England/ALDP-C3AFB9/2020\|EPI_ISL_731796\|20 | .................... | .................... | .................... | .................... | .................... | .................... | |
| Y | 7168 | hCoV-19/England/ALDP-C3BF9A/2020\|EPI_ISL_731798\|20 | .................... | .................... | .................... | .................... | .................... | .................... | |
| Y | 7169 | hCoV-19/England/LOND-DA5E2/2020\|EPI_ISL_724645\|202 | ..................T. | .................... | .................... | .................... | .................... | .................... | 1 |
| Y | 7170 | hCoV-19/England/LOND-DA649/2020\|EPI_ISL_724649\|202 | .................... | .A.................. | .................... | .................... | .................... | ..A................. | |
| Y | 7171 | hCoV-19/England/LOND-DA667/2020\|EPI_ISL_724651\|202 | .................... | .................... | .................... | .................... | .................... | .................... | |
| Y | 7172 | hCoV-19/England/LOND-DA6D0/2020\|EPI_ISL_724655\|202 | .................... | .A.................. | .................... | .................... | .................... | ..A................. | |
| Y | 7173 | hCoV-19/England/LOND-DA6EF/2020\|EPI_ISL_724656\|202 | .................... | .A.................. | .................... | .................... | .................... | ..A................. | |
| Y | 7174 | hCoV-19/England/LOND-DA728/2020\|EPI_ISL_724659\|202 | .................... | .A.................. | .................... | .................... | .................... | ..A................. | |
| Y | 7175 | hCoV-19/England/LOND-DA755/2020\|EPI_ISL_724661\|202 | .................... | .................... | .................... | .................... | .................... | .................... | |
| Y | 7176 | hCoV-19/England/LOND-DA773/2020\|EPI_ISL_724663\|202 | .................... | .................... | .................... | .................... | .................... | .................... | |
| Y | 7177 | hCoV-19/England/LOND-DA807/2020\|EPI_ISL_724664\|202 | .................... | .A.................. | .................... | .................... | .................... | ..A................. | |
| Y | 7178 | hCoV-19/England/LOND-DA8CB/2020\|EPI_ISL_724668\|202 | .................... | .................... | .................... | .................... | .................... | .................... | |
| Y | 7179 | hCoV-19/England/LOND-DAB3B/2020\|EPI_ISL_724695\|202 | .................... | .................... | .................... | .................... | .................... | .................... | |
| Y | 7180 | hCoV-19/England/LOND-DAB86/2020\|EPI_ISL_724699\|202 | .................... | .................... | .................... | .................... | ....T............... | .................... | |
| Y | 7181 | hCoV-19/Wales/PHWC-49360C/2020\|EPI_ISL_725915\|2020 | .................... | .................... | .................... | .................... | .................... | .................... | |
| Y | 7182 | hCoV-19/Wales/PHWC-49361B/2020\|EPI_ISL_725916\|2020 | .................... | .................... | .................... | .................... | .................... | .................... | |
| Y | 7183 | hCoV-19/Wales/PHWC-49362A/2020\|EPI_ISL_725917\|2020 | .................... | .................... | .................... | .................... | .................... | .................... | |
| Y | 7184 | hCoV-19/Wales/PHWC-493657/2020\|EPI_ISL_725919\|2020 | .................... | .................... | .................... | .................... | .................... | .................... | |
| Y | 7185 | hCoV-19/Wales/PHWC-493666/2020\|EPI_ISL_725920\|2020 | .................... | .................... | .................... | .................... | .................... | .................... | |
| Y | 7186 | hCoV-19/Wales/PHWC-493675/2020\|EPI_ISL_725921\|2020 | .................... | .................... | .................... | .................... | .................... | .................... | |
| Y | 7188 | hCoV-19/Wales/PHWC-4936A2/2020\|EPI_ISL_725924\|2020 | .................... | .................... | .................... | .................... | .................... | .................... | |
| Y | 7189 | hCoV-19/Wales/PHWC-4936B1/2020\|EPI_ISL_725925\|2020 | .................... | .................... | .................... | .................... | .................... | .................... | |
| Y | 7190 | hCoV-19/Wales/PHWC-4936C0/2020\|EPI_ISL_725926\|2020 | .................... | .................... | .................... | .................... | .................... | .................... | |
| Y | 7191 | hCoV-19/Wales/PHWC-4936DF/2020\|EPI_ISL_725927\|2020 | .................... | .................... | .................... | .................... | .................... | .................... | |
| Y | 7192 | hCoV-19/Wales/PHWC-4936EE/2020\|EPI_ISL_725928\|2020 | .................... | .................... | .................... | .................... | .................... | .................... | |
| Y | 7193 | hCoV-19/Wales/PHWC-4936FD/2020\|EPI_ISL_725929\|2020 | .................... | .................... | .................... | .................... | .................... | .................... | |
| Y | 7194 | hCoV-19/Wales/PHWC-493709/2020\|EPI_ISL_725930\|2020 | .................... | .................... | .................... | .................... | .................... | .................... | |
| Y | 7195 | hCoV-19/Wales/PHWC-493727/2020\|EPI_ISL_725932\|2020 | .................... | .................... | .................... | .................... | .................... | .................... | |
| Y | 7196 | hCoV-19/Wales/PHWC-493736/2020\|EPI_ISL_725933\|2020 | .................... | .................... | .................... | .................... | .................... | .................... | |
| Y | 7197 | hCoV-19/Wales/PHWC-493745/2020\|EPI_ISL_725934\|2020 | .................... | .................... | .................... | .................... | .................... | .................... | |
| Y | 7198 | hCoV-19/Wales/PHWC-493754/2020\|EPI_ISL_725935\|2020 | .................... | .................... | .................... | .................... | .................... | .................... | |
| Y | 7199 | hCoV-19/Wales/PHWC-493763/2020\|EPI_ISL_725936\|2020 | .................... | .................... | .................... | .................... | .................... | .................... | |
| Y | 7200 | hCoV-19/Wales/PHWC-493772/2020\|EPI_ISL_725937\|2020 | .................... | .................... | .................... | .................... | .................... | .................... | |
| Y | 7201 | hCoV-19/Wales/PHWC-493781/2020\|EPI_ISL_725938\|2020 | .................... | .................... | .................... | .................... | .................... | .................... | |
| Y | 7202 | hCoV-19/Wales/PHWC-493790/2020\|EPI_ISL_725939\|2020 | .................... | .................... | .................... | .................... | .................... | .................... | |
| Y | 7203 | hCoV-19/Wales/PHWC-4937AF/2020\|EPI_ISL_725940\|2020 | .................... | .................... | .................... | .................... | .................... | .................... | |
| Y | 7204 | hCoV-19/Wales/PHWC-4937BE/2020\|EPI_ISL_725941\|2020 | .................... | .................... | .................... | .................... | .................... | .................... | |
| Y | 7205 | hCoV-19/Wales/PHWC-4937CD/2020\|EPI_ISL_725942\|2020 | .................... | .................... | .................... | .................... | .................... | .................... | |
| Y | 7206 | hCoV-19/Wales/PHWC-4937DC/2020\|EPI_ISL_725943\|2020 | .................... | .................... | .................... | .................... | .................... | .................... | |
| Y | 7207 | hCoV-19/Wales/PHWC-4937EB/2020\|EPI_ISL_725944\|2020 | .................... | .................... | .................... | .................... | .................... | .................... | |
| Y | 7208 | hCoV-19/Wales/PHWC-4937FA/2020\|EPI_ISL_725945\|2020 | .................... | .................... | .................... | .................... | .................... | .................... | |
| Y | 7209 | hCoV-19/Wales/PHWC-493806/2020\|EPI_ISL_725946\|2020 | .................... | .................... | .................... | .................... | .................... | .................... | |
| Y | 7210 | hCoV-19/Wales/PHWC-493815/2020\|EPI_ISL_725947\|2020 | .................... | .................... | .................... | .................... | .................... | .................... | |
| Y | 7211 | hCoV-19/Wales/PHWC-493824/2020\|EPI_ISL_725948\|2020 | .................... | .................... | .................... | .................... | .................... | .................... | |
| Y | 7212 | hCoV-19/Wales/PHWC-493842/2020\|EPI_ISL_725949\|2020 | .................... | .................... | .................... | .................... | .................... | .................... | |
| Y | 7213 | hCoV-19/Wales/PHWC-493860/2020\|EPI_ISL_725950\|2020 | .................... | .................... | .................... | .................... | .................... | .................... | |
| Y | 7215 | hCoV-19/Wales/PHWC-4938AC/2020\|EPI_ISL_725952\|2020 | .................... | .................... | .................... | .................... | .................... | .................... | |
| Y | 7216 | hCoV-19/Wales/PHWC-4938BB/2020\|EPI_ISL_725953\|2020 | .................... | .................... | .................... | .................... | .................... | .................... | |
| Y | 7217 | hCoV-19/Wales/PHWC-4938CA/2020\|EPI_ISL_725954\|2020 | .................... | .................... | .................... | .................... | .................... | .................... | |
| Y | 7218 | hCoV-19/Wales/PHWC-4938E8/2020\|EPI_ISL_725955\|2020 | .................... | .................... | .................... | .................... | .................... | .................... | |
| Y | 7221 | hCoV-19/Wales/PHWC-493912/2020\|EPI_ISL_725958\|2020 | .................... | .................... | .................... | .................... | .................... | .................... | |
| Y | 7222 | hCoV-19/Wales/PHWC-49394F/2020\|EPI_ISL_725959\|2020 | .................... | .................... | .................... | .................... | .................... | .................... | |
| Y | 7224 | hCoV-19/Wales/PHWC-49397C/2020\|EPI_ISL_725962\|2020 | .................... | .................... | .................... | .................... | .................... | .................... | |
| Y | 7226 | hCoV-19/Wales/PHWC-49399A/2020\|EPI_ISL_725964\|2020 | .................... | .................... | .................... | .................... | .................... | .................... | |
| Y | 7227 | hCoV-19/Wales/PHWC-4939A9/2020\|EPI_ISL_725965\|2020 | .................... | .................... | .................... | .................... | .................... | .................... | |
| Y | 7228 | hCoV-19/Wales/PHWC-4939C7/2020\|EPI_ISL_725967\|2020 | .................... | .....A.............. | .................... | .................... | .................... | .................... | |
| Y | 7229 | hCoV-19/Wales/PHWC-4939D6/2020\|EPI_ISL_725968\|2020 | .................... | .................... | .................... | .................... | .................... | .................... | |
| Y | 7230 | hCoV-19/Wales/PHWC-4939E5/2020\|EPI_ISL_725969\|2020 | .................... | .................... | .................... | .................... | .................... | .................... | |
| Y | 7231 | hCoV-19/Wales/PHWC-493A00/2020\|EPI_ISL_725971\|2020 | .................... | .................... | .................... | .................... | .................... | .................... | |
| Y | 7232 | hCoV-19/Wales/PHWC-493A1F/2020\|EPI_ISL_725972\|2020 | .................... | .................... | .................... | .................... | .................... | .................... | |
| Y | 7234 | hCoV-19/Wales/PHWC-493A3D/2020\|EPI_ISL_725974\|2020 | .................... | .................... | .................... | .................... | .................... | .................... | |
| Y | 7235 | hCoV-19/Wales/PHWC-493A4C/2020\|EPI_ISL_725975\|2020 | .................... | .................... | .................... | .................... | .................... | .................... | |
| Y | 7236 | hCoV-19/Wales/PHWC-493A5B/2020\|EPI_ISL_725976\|2020 | .................... | .....A.............. | .................... | .................... | .................... | .................... | |
| Y | 7237 | hCoV-19/Wales/PHWC-493A6A/2020\|EPI_ISL_725977\|2020 | .................... | .................... | .................... | .................... | .................... | .................... | |
| Y | 7239 | hCoV-19/Wales/PHWC-493A88/2020\|EPI_ISL_725979\|2020 | .................... | .................... | .................... | .................... | .................... | .................... | |
| Y | 7240 | hCoV-19/Wales/PHWC-493A97/2020\|EPI_ISL_725980\|2020 | .................... | .................... | .................... | .................... | .................... | .................... | |
| Y | 7241 | hCoV-19/Wales/PHWC-493AB5/2020\|EPI_ISL_725982\|2020 | .................... | .................... | .................... | .................... | .................... | .................... | |
| Y | 7242 | hCoV-19/Wales/PHWC-493AD3/2020\|EPI_ISL_725984\|2020 | .................... | .....A.............. | .................... | .................... | .................... | .................... | |
| Y | 7243 | hCoV-19/Wales/PHWC-493B1C/2020\|EPI_ISL_725986\|2020 | .................... | .................... | .................... | .................... | .................... | .................... | |
| Y | 7244 | hCoV-19/Wales/PHWC-493B2B/2020\|EPI_ISL_725987\|2020 | .................... | .................... | .................... | .................... | .................... | .................... | |
| Y | 7245 | hCoV-19/Wales/PHWC-493B3A/2020\|EPI_ISL_725988\|2020 | .................... | .................... | .................... | .................... | .................... | .................... | |
| Y | 7246 | hCoV-19/Wales/PHWC-493B85/2020\|EPI_ISL_725991\|2020 | .................... | .................... | .................... | .................... | .................... | .................... | |
| Y | 7247 | hCoV-19/Wales/PHWC-493BA3/2020\|EPI_ISL_725992\|2020 | .................... | .................... | .................... | .................... | .................... | .................... | |
| Y | 7248 | hCoV-19/Wales/PHWC-493BB2/2020\|EPI_ISL_725993\|2020 | .................... | .................... | .................... | .................... | .................... | .................... | |
| Y | 7250 | hCoV-19/Wales/PHWC-493BEF/2020\|EPI_ISL_725996\|2020 | .................... | .................... | .................... | .................... | .................... | .................... | |
| Y | 7251 | hCoV-19/Wales/PHWC-493BFE/2020\|EPI_ISL_725997\|2020 | .................... | .................... | .................... | .................... | .................... | .................... | |
| Y | 7252 | hCoV-19/Wales/PHWC-493C19/2020\|EPI_ISL_725999\|2020 | .................... | .................... | .................... | .................... | .................... | .................... | |

| | | | | | | | | |
|---|---|---|---|---|---|---|---|---|
| Y | 7253 | hCoV-19/Malaysia/IMR-WC194899/2020\|EPI_ISL_728160\| | ···················· | ·A·················· | ···················· | ···················· | ···················· | ···················· |
| Y | 7254 | hCoV-19/Malaysia/IMR-WC196746/2020\|EPI_ISL_728161\| | ···················· | ·A·················· | ···················· | ···················· | ···················· | ···················· |
| Y | 7255 | hCoV-19/Singapore/1454/2020\|EPI_ISL_728190\|2020-12 | ···················· | ···················· | ···················· | ···················· | ···················· | ··A················· |
| Y | 7256 | hCoV-19/Singapore/1455/2020\|EPI_ISL_728191\|2020-12 | ···················· | ···················· | ···················· | ···················· | ···················· | ···················· |
| Y | 7257 | hCoV-19/Singapore/1456/2020\|EPI_ISL_728192\|2020-12 | ···················· | ···················· | ···················· | ···················· | ···················· | ···················· |
| Y | 7258 | hCoV-19/Singapore/1457/2020\|EPI_ISL_728193\|2020-12 | ···················· | ···················· | ···················· | ···················· | ···················· | ··A················· |
| Y | 7259 | hCoV-19/Singapore/1458/2020\|EPI_ISL_728194\|2020-12 | ···················· | ·A·················· | ···················· | ···················· | ···················· | ···················· |
| Y | 7260 | hCoV-19/Singapore/1459/2020\|EPI_ISL_728195\|2020-12 | ···················· | ·A·················· | ···················· | ···················· | ···················· | ···················· |
| Y | 7261 | hCoV-19/Singapore/1460/2020\|EPI_ISL_728196\|2020-12 | ···················· | ···················· | ···················· | ···················· | ···················· | ···················· |
| Y | 7262 | hCoV-19/Singapore/1462/2020\|EPI_ISL_728198\|2020-12 | ···················· | ···················· | ···················· | ···················· | ···················· | ···················· |
| Y | 7263 | hCoV-19/Singapore/1463/2020\|EPI_ISL_728199\|2020-12 | ···················· | ·A·················· | ···················· | ···················· | ···················· | ··A················· |
| Y | 7264 | hCoV-19/Germany/DE-BE-ChVir1900/2020\|EPI_ISL_72948 | ···················· | ···················· | ···················· | ···················· | ···················· | ···················· |
| Y | 7265 | hCoV-19/Germany/DE-BW-ChVir1902/2020\|EPI_ISL_72948 | ···················· | ···················· | ···················· | ···················· | ···················· | ···················· |
| Y | 7266 | hCoV-19/Germany/DE-RP-ChVir1906/2020\|EPI_ISL_72948 | ···················· | ···················· | ···················· | ···················· | ···················· | ···················· |
| Y | 7267 | hCoV-19/Germany/DE-RP-ChVir1908/2020\|EPI_ISL_72948 | ···················· | ···················· | ···················· | ···················· | ···················· | ···················· |
| Y | 7268 | hCoV-19/Germany/DE-BE-ChVir1919/2020\|EPI_ISL_72948 | ···················· | ···················· | ···················· | ···················· | ···················· | ···················· |
| Y | 7269 | hCoV-19/Germany/DE-BW-ChVir1946/2020\|EPI_ISL_72948 | ···················· | ···················· | ···················· | ···················· | ···················· | ···················· |
| Y | 7270 | hCoV-19/Germany/DE-BE-ChVir1959/2020\|EPI_ISL_72948 | ···················· | ···················· | ···················· | ···················· | ···················· | ···················· |
| Y | 7271 | hCoV-19/Jordan/IR-ALSR-5232/2020\|EPI_ISL_730470\|20 | ···················· | ···················· | ···················· | ···················· | ···················· | ···················· |
| Y | 7272 | hCoV-19/Jordan/IR-ALSR-5233/2020\|EPI_ISL_730471\|20 | ···················· | ·A·················· | ···················· | ···················· | ···················· | ··A················· |
| Y | 7273 | hCoV-19/Jordan/IR-ALSR-5234/2020\|EPI_ISL_730472\|20 | ···················· | ·A·················· | ···················· | ···················· | ···················· | ··A················· |
| Y | 7274 | hCoV-19/Jordan/IR-ALSR-5236/2020\|EPI_ISL_730474\|20 | ···················· | ···················· | ···················· | ···················· | ···················· | ···················· |
| Y | 7275 | hCoV-19/Jordan/IR-ALSR-5237/2020\|EPI_ISL_730475\|20 | ···················· | ···················· | ···················· | ···················· | ···················· | ···················· |
| Y | 7276 | hCoV-19/Jordan/IR-ALSR-5238/2020\|EPI_ISL_730476\|20 | ···················· | ···················· | ···················· | ···················· | ···················· | ···················· |
| Y | 7277 | hCoV-19/Jordan/IR-ALSR-5239/2020\|EPI_ISL_730477\|20 | ···················· | ···················· | ···················· | ···················· | ···················· | ···················· |
| Y | 7278 | hCoV-19/Jordan/AQ-ALSR-5240/2020\|EPI_ISL_730478\|20 | ···················· | ···················· | ···················· | ···················· | ···················· | ···················· |
| Y | 7279 | hCoV-19/England/QEUH-C33EE0/2020\|EPI_ISL_734017\|20 | ···················· | ···················· | ···················· | ···················· | ···················· | ···················· |
| Y | 7280 | hCoV-19/England/QEUH-C34016/2020\|EPI_ISL_734018\|20 | ···················· | ···················· | ···················· | ···················· | ···················· | ···················· |
| Y | 7281 | hCoV-19/England/QEUH-C342D4/2020\|EPI_ISL_734019\|20 | ···················· | ···················· | ···················· | ···················· | ···················· | ···················· |
| Y | 7282 | hCoV-19/England/QEUH-C34113/2020\|EPI_ISL_734020\|20 | ···················· | ···················· | ···················· | ···················· | ···················· | ···················· |
| Y | 7283 | hCoV-19/England/QEUH-C344B0/2020\|EPI_ISL_734021\|20 | ···················· | ···················· | ···················· | ···················· | ···················· | ···················· |
| Y | 7284 | hCoV-19/England/QEUH-C33BDA/2020\|EPI_ISL_734022\|20 | ···················· | ···················· | ···················· | ···················· | ···················· | ···················· |
| Y | 7285 | hCoV-19/England/QEUH-C33A0A/2020\|EPI_ISL_734023\|20 | ···················· | ···················· | ···················· | ···················· | ···················· | ···················· |
| Y | 7286 | hCoV-19/England/QEUH-C347E4/2020\|EPI_ISL_734024\|20 | ···················· | ···················· | ···················· | ···················· | ···················· | ···················· |
| Y | 7287 | hCoV-19/England/QEUH-C3393A/2020\|EPI_ISL_734025\|20 | ···················· | ···················· | ···················· | ···················· | ···················· | ···················· |
| Y | 7288 | hCoV-19/England/QEUH-C34438/2020\|EPI_ISL_734027\|20 | ···················· | ···················· | ···················· | ···················· | ···················· | ···················· |
| Y | 7289 | hCoV-19/England/QEUH-C338E2/2020\|EPI_ISL_734029\|20 | ···················· | ···················· | ···················· | ···················· | ···················· | ···················· |
| Y | 7290 | hCoV-19/Turkey/IST-GLAB-233/2020\|EPI_ISL_735300\|20 | ···················· | ···················· | ···················· | ···················· | ···················· | ···················· |
| Y | 7291 | hCoV-19/Turkey/IST-GLAB-234/2020\|EPI_ISL_735301\|20 | ···················· | ···················· | ···················· | ···················· | ···················· | ···················· |
| Y | 7292 | hCoV-19/Turkey/IST-GLAB-235/2020\|EPI_ISL_735302\|20 | ···················· | ···················· | ···················· | ···················· | ···················· | ···················· |
| Y | 7293 | hCoV-19/Turkey/IST-GLAB-236/2020\|EPI_ISL_735303\|20 | ···················· | ···················· | ···················· | ···················· | ···················· | ···················· |
| Y | 7294 | hCoV-19/Turkey/IST-GLAB-237/2020\|EPI_ISL_735304\|20 | ···················· | ···················· | ···················· | ···················· | ···················· | ···················· |
| Y | 7295 | hCoV-19/Turkey/IST-GLAB-238/2020\|EPI_ISL_735305\|20 | ···················· | ···················· | ···················· | ···················· | ···················· | ···················· |
| Y | 7296 | hCoV-19/Turkey/IST-GLAB-239/2020\|EPI_ISL_735306\|20 | ···················· | ···················· | ···················· | ···················· | ···················· | ···················· |
| Y | 7297 | hCoV-19/Turkey/IST-GLAB-240/2020\|EPI_ISL_735307\|20 | ···················· | ···················· | ···················· | ···················· | ···················· | ···················· |
| Y | 7298 | hCoV-19/Turkey/IST-GLAB-241/2020\|EPI_ISL_735308\|20 | ···················· | ···················· | ···················· | ···················· | ···················· | ···················· |
| Y | 7299 | hCoV-19/Turkey/IST-GLAB-243/2020\|EPI_ISL_735309\|20 | ···················· | ···················· | ···················· | ···················· | ···················· | ···················· |
| Y | 7300 | hCoV-19/Turkey/IST-GLAB-244/2020\|EPI_ISL_735310\|20 | ···················· | ···················· | ···················· | ···················· | ···················· | ···················· |
| Y | 7301 | hCoV-19/Turkey/IST-GLAB-245/2020\|EPI_ISL_735311\|20 | ···················· | ···················· | ···················· | ···················· | ···················· | ···················· |
| Y | 7302 | hCoV-19/Turkey/IST-GLAB-246/2020\|EPI_ISL_735312\|20 | ···················· | ···················· | ···················· | ···················· | ···················· | ···················· |
| Y | 7303 | hCoV-19/Turkey/IST-GLAB-247/2020\|EPI_ISL_735313\|20 | ···················· | ···················· | ···················· | ···················· | ···················· | ···················· |
| Y | 7304 | hCoV-19/Turkey/IST-GLAB-248/2020\|EPI_ISL_735314\|20 | ···················· | ···················· | ···················· | ···················· | ···················· | ···················· |
| Y | 7305 | hCoV-19/Turkey/IST-GLAB-249/2020\|EPI_ISL_735315\|20 | ···················· | ···················· | ···················· | ···················· | ···················· | ···················· |
| Y | 7306 | hCoV-19/Turkey/IST-GLAB-250/2020\|EPI_ISL_735316\|20 | ···················· | ···················· | ···················· | ···················· | ···················· | ···················· |
| Y | 7307 | hCoV-19/Turkey/IST-GLAB-252/2020\|EPI_ISL_735317\|20 | ···················· | ···················· | ···················· | ···················· | ···················· | ···················· |
| Y | 7308 | hCoV-19/Turkey/IST-GLAB-253/2020\|EPI_ISL_735318\|20 | ···················· | ···················· | ···················· | ···················· | ···················· | ···················· |
| Y | 7309 | hCoV-19/Turkey/IST-GLAB-304/2020\|EPI_ISL_735344\|20 | ···················· | ···················· | ···················· | ···················· | ···················· | ···················· |
| Y | 7310 | hCoV-19/Turkey/IST-GLAB-311/2020\|EPI_ISL_735345\|20 | ···················· | ···················· | ···················· | ···················· | ···················· | ···················· |
| Y | 7311 | hCoV-19/Turkey/IST-GLAB-319/2020\|EPI_ISL_735348\|20 | ···················· | ···················· | ···················· | ···················· | ···················· | ···················· |
| Y | 7312 | hCoV-19/Turkey/IST-GLAB-320/2020\|EPI_ISL_735349\|20 | ···················· | ···················· | ···················· | ···················· | ···················· | ···················· |
| Y | 7313 | hCoV-19/Turkey/IST-GLAB-323/2020\|EPI_ISL_735350\|20 | ···················· | ···················· | ···················· | ···················· | ···················· | ···················· |
| Y | 7314 | hCoV-19/Turkey/IST-GLAB-325/2020\|EPI_ISL_735351\|20 | ···················· | ···················· | ···················· | ···················· | ···················· | ···················· |
| Y | 7315 | hCoV-19/Turkey/IST-GLAB-335/2020\|EPI_ISL_735352\|20 | ···················· | ···················· | ···················· | ···················· | ···················· | ···················· |
| Y | 7316 | hCoV-19/Turkey/IST-GLAB-337/2020\|EPI_ISL_735353\|20 | ···················· | ···················· | ···················· | ···················· | ···················· | ···················· |
| Y | 7317 | hCoV-19/Turkey/IST-GLAB-338/2020\|EPI_ISL_735354\|20 | ···················· | ···················· | ···················· | ···················· | ···················· | ···················· |
| Y | 7318 | hCoV-19/Turkey/IST-GLAB-340/2020\|EPI_ISL_735355\|20 | ···················· | ···················· | ···················· | ···················· | ···················· | ···················· |
| Y | 7319 | hCoV-19/Turkey/IST-GLAB-341/2020\|EPI_ISL_735356\|20 | ···················· | ···················· | ···················· | ···················· | ···················· | ···················· |
| Y | 7320 | hCoV-19/Turkey/IST-GLAB-342/2020\|EPI_ISL_735357\|20 | ···················· | ···················· | ···················· | ···················· | ···················· | ···················· |
| Y | 7321 | hCoV-19/Turkey/IST-GLAB-343/2020\|EPI_ISL_735358\|20 | ···················· | ···················· | ···················· | ···················· | ···················· | ···················· |
| Y | 7322 | hCoV-19/Turkey/IST-GLAB-347/2020\|EPI_ISL_735359\|20 | ···················· | ···················· | ···················· | ···················· | ···················· | ···················· |
| Y | 7323 | hCoV-19/England/ALDP-C48D72/2020\|EPI_ISL_736600\|20 | ···················· | ···················· | ···················· | ···················· | ···················· | ···················· |
| Y | 7324 | hCoV-19/England/ALDP-C4847D/2020\|EPI_ISL_736604\|20 | ···················· | ···················· | ···················· | ···················· | ···················· | ···················· |
| Y | 7325 | hCoV-19/England/QEUH-C4BB57/2020\|EPI_ISL_736605\|20 | ···················· | ···················· | ···················· | ···················· | ···················· | ···················· |
| Y | 7326 | hCoV-19/England/ALDP-C487DE/2020\|EPI_ISL_736607\|20 | ···················· | ···················· | ···················· | ···················· | ···················· | ···················· |
| Y | 7327 | hCoV-19/England/MILK-C50876/2020\|EPI_ISL_736610\|20 | ···················· | ···················· | ···················· | ···················· | ······T············· | ···················· |
| Y | 7328 | hCoV-19/England/ALDP-C48A8A/2020\|EPI_ISL_736612\|20 | ···················· | ···················· | ···················· | ···················· | ···················· | ···················· |
| Y | 7329 | hCoV-19/England/ALDP-C48D36/2020\|EPI_ISL_736613\|20 | ···················· | ·A·················· | ···················· | ···················· | ···················· | ··A················· |
| Y | 7330 | hCoV-19/England/ALDP-C489F6/2020\|EPI_ISL_736614\|20 | ···················· | ···················· | ···················· | ···················· | ···················· | ···················· |
| Y | 7331 | hCoV-19/England/ALDP-C4861D/2020\|EPI_ISL_736615\|20 | ···········T········ | ···················· | ···················· | ···················· | ···················· | ···················· |
| Y | 7332 | hCoV-19/England/ALDP-C48DFA/2020\|EPI_ISL_736616\|20 | ···················· | ···················· | ···················· | ···················· | ···················· | ···················· |

| | | | | | | | |
|---|---|---|---|---|---|---|---|
| Y | 7333 hCoV-19/England/MILK-C50885/2020\|EPI_ISL_736617\|20 | ···················· | ···················· | ···················· | ···················· | ······T············· | ···················· |
| Y | 7334 hCoV-19/England/ALDP-C48C93/2020\|EPI_ISL_736621\|20 | ···················· | ···················· | ···················· | ···················· | ···················· | ···················· |
| Y | 7335 hCoV-19/England/QEUH-C4BCDC/2020\|EPI_ISL_736624\|20 | ···················· | ···················· | ···················· | ···················· | ···················· | ···················· |
| Y | 7336 hCoV-19/England/ALDP-C48DAF/2020\|EPI_ISL_736626\|20 | ···················· | ···················· | ···················· | ···················· | ···················· | ···················· |
| Y | 7337 hCoV-19/England/QEUH-C4BD7F/2020\|EPI_ISL_736627\|20 | ···················· | ···················· | ···················· | ···················· | ···················· | ···················· |
| Y | 7338 hCoV-19/England/QEUH-C4BCEB/2020\|EPI_ISL_736628\|20 | ···················· | ···················· | ···················· | ···················· | ···················· | ···················· |
| Y | 7339 hCoV-19/England/QEUH-C4BD15/2020\|EPI_ISL_736631\|20 | ···················· | ···················· | ···················· | ···················· | ···················· | ···················· |
| Y | 7340 hCoV-19/England/ALDP-C4846E/2020\|EPI_ISL_736634\|20 | ···················· | ···················· | ···················· | ···················· | ···················· | ···················· |
| Y | 7341 hCoV-19/England/MILK-C5064F/2020\|EPI_ISL_736637\|20 | ···················· | ···················· | ···················· | ···················· | ···················· | ···················· |
| Y | 7342 hCoV-19/England/ALDP-C48A20/2020\|EPI_ISL_736639\|20 | ···················· | ·A·················· | ···················· | ···················· | ···················· | ···················· |
| Y | 7343 hCoV-19/England/ALDP-C4856B/2020\|EPI_ISL_736641\|20 | ···················· | ···················· | ···················· | ···················· | ······T············· | ···················· |
| Y | 7344 hCoV-19/England/ALDP-C48307/2020\|EPI_ISL_736642\|20 | ···················· | ···················· | ···················· | ···················· | ···················· | ···················· |
| Y | 7345 hCoV-19/England/ALDP-C48AE4/2020\|EPI_ISL_736644\|20 | ···················· | ···················· | ···················· | ···················· | ······T············· | ···················· |
| Y | 7346 hCoV-19/England/QEUH-C4BDCA/2020\|EPI_ISL_736646\|20 | ···················· | ···················· | ···················· | ···················· | ···················· | ···················· |
| Y | 7347 hCoV-19/England/ALDP-C48BE1/2020\|EPI_ISL_736647\|20 | ···················· | ···················· | ···················· | ···················· | ···················· | ···················· |
| Y | 7348 hCoV-19/England/ALDP-C4845F/2020\|EPI_ISL_736649\|20 | ···················· | ···················· | ···················· | ···················· | ···················· | ···················· |
| Y | 7349 hCoV-19/England/ALDP-C48B69/2020\|EPI_ISL_736680\|20 | ···················· | ···················· | ···················· | ···················· | ···················· | ···················· |
| Y | 7350 hCoV-19/England/MILK-C507C4/2020\|EPI_ISL_736682\|20 | ···················· | ·A·················· | ···················· | ···················· | ···················· | ··A················· |
| Y | 7351 hCoV-19/England/ALDP-C48659/2020\|EPI_ISL_736687\|20 | ···················· | ···················· | ···················· | ···················· | ···················· | ···················· |
| Y | 7352 hCoV-19/England/ALDP-C48756/2020\|EPI_ISL_736688\|20 | ···················· | ···················· | ···················· | ···················· | ···················· | ···················· |
| Y | 7353 hCoV-19/Switzerland/VD-ETHZ-320587/2020\|EPI_ISL_73 | ···················· | ···················· | ···················· | ···················· | ···················· | ···················· |
| Y | 7354 hCoV-19/Switzerland/VD-ETHZ-410088/2020\|EPI_ISL_73 | ···················· | ···················· | ···················· | ···················· | ···················· | ···················· |
| Y | 7355 hCoV-19/Switzerland/VD-ETHZ-410190/2020\|EPI_ISL_73 | ···················· | ···················· | ···················· | ···················· | ···················· | ···················· |
| Y | 7356 hCoV-19/Switzerland/VD-ETHZ-410552/2020\|EPI_ISL_73 | ···················· | ···················· | ···················· | ···················· | ···················· | ···················· |
| Y | 7357 hCoV-19/Switzerland/VD-ETHZ-410553/2020\|EPI_ISL_73 | ···················· | ···················· | ···················· | ···················· | ···················· | ···················· |
| Y | 7358 hCoV-19/Switzerland/VD-ETHZ-410554/2020\|EPI_ISL_73 | ···················· | ···················· | ···················· | ···················· | ···················· | ···················· |
| Y | 7359 hCoV-19/Switzerland/VD-ETHZ-410555/2020\|EPI_ISL_73 | ···················· | ···················· | ···················· | ···················· | ···················· | ···················· |
| Y | 7360 hCoV-19/Switzerland/VD-ETHZ-410924/2020\|EPI_ISL_73 | ···················· | ···················· | ···················· | ···················· | ···················· | ···················· |
| Y | 7361 hCoV-19/Switzerland/ZG-ETHZ-410276/2020\|EPI_ISL_73 | ···················· | ···················· | ···················· | ···················· | ···················· | ···················· |
| Y | 7362 hCoV-19/Switzerland/ZH-ETHZ-410024/2020\|EPI_ISL_73 | ···················· | ···················· | ···················· | ···················· | ···················· | ···················· |
| Y | 7363 hCoV-19/Switzerland/ZH-ETHZ-410025/2020\|EPI_ISL_73 | ···················· | ···················· | ···················· | ···················· | ···················· | ···················· |
| Y | 7364 hCoV-19/Switzerland/ZH-ETHZ-410026/2020\|EPI_ISL_73 | ···················· | ···················· | ···················· | ···················· | ···················· | ···················· |
| Y | 7365 hCoV-19/Switzerland/ZH-ETHZ-410046/2020\|EPI_ISL_73 | ···················· | ···················· | ···················· | ···················· | ···················· | ···················· |
| Y | 7366 hCoV-19/Switzerland/ZH-ETHZ-410064/2020\|EPI_ISL_73 | ···················· | ···················· | ···················· | ···················· | ···················· | ···················· |
| Y | 7367 hCoV-19/Switzerland/ZH-ETHZ-410080/2020\|EPI_ISL_73 | ···················· | ···················· | ···················· | ···················· | ···················· | ···················· |
| Y | 7368 hCoV-19/Switzerland/ZH-ETHZ-410093/2020\|EPI_ISL_73 | ···················· | ···················· | ···················· | ···················· | ···················· | ···················· |
| Y | 7369 hCoV-19/Switzerland/ZH-ETHZ-410094/2020\|EPI_ISL_73 | ···················· | ···················· | ···················· | ···················· | ···S················ | ···················· |
| Y | 7370 hCoV-19/Switzerland/ZH-ETHZ-410165/2020\|EPI_ISL_73 | ···················· | ···················· | ···················· | ···················· | ···················· | ···················· |
| Y | 7371 hCoV-19/Switzerland/ZH-ETHZ-410224/2020\|EPI_ISL_73 | ···················· | ···················· | ···················· | ···················· | ···················· | ···················· |
| Y | 7373 hCoV-19/Switzerland/ZH-ETHZ-410529/2020\|EPI_ISL_73 | ···················· | ···················· | ···················· | ···················· | ···················· | ···················· |
| Y | 7374 hCoV-19/Switzerland/ZH-ETHZ-410534/2020\|EPI_ISL_73 | ···················· | ···················· | ···················· | ···················· | ···················· | ···················· |
| Y | 7375 hCoV-19/Switzerland/ZH-ETHZ-410541/2020\|EPI_ISL_73 | ···················· | ···················· | ···················· | ···················· | ···················· | ···················· |
| Y | 7376 hCoV-19/Switzerland/ZH-ETHZ-410542/2020\|EPI_ISL_73 | ···················· | ···················· | ···················· | ···················· | ···················· | ···················· |
| Y | 7377 hCoV-19/Switzerland/ZH-ETHZ-410551/2020\|EPI_ISL_73 | ···················· | ···················· | ···················· | ···················· | ···················· | ···················· |
| Y | 7378 hCoV-19/Switzerland/ZH-ETHZ-410896/2020\|EPI_ISL_73 | ···················· | ········A··········· | ···················· | ···················· | ···················· | ···················· |
| Y | 7379 hCoV-19/Switzerland/ZH-ETHZ-410897/2020\|EPI_ISL_73 | ···················· | ········A··········· | ···················· | ···················· | ···················· | ···················· |
| Y | 7380 hCoV-19/Switzerland/ZH-ETHZ-410898/2020\|EPI_ISL_73 | ···················· | ···················· | ···················· | ···················· | ······K············· | ···················· |
| Y | 7381 hCoV-19/Switzerland/ZH-ETHZ-410904/2020\|EPI_ISL_73 | ···················· | ···················· | ···················· | ···················· | ···················· | ···················· |
| Y | 7382 hCoV-19/Switzerland/ZH-ETHZ-410911/2020\|EPI_ISL_73 | ···················· | ···················· | ···················· | ···················· | ···················· | ···················· |
| Y | 7383 hCoV-19/Uganda/UG047/2020\|EPI_ISL_737935\|2020-04-0 | ···················· | ···················· | ···················· | ···················· | ···················· | ···················· |
| Y | 7384 hCoV-19/Uganda/UG046/2020\|EPI_ISL_737939\|2020-07-1 | ···················· | ···················· | ···················· | ···················· | ···················· | ···················· |
| Y | 7385 hCoV-19/Uganda/UG066/2020\|EPI_ISL_737940\|2020-08-2 | ···················· | ···················· | ···················· | ···················· | ···················· | ···················· |
| Y | 7386 hCoV-19/Uganda/UG068/2020\|EPI_ISL_737941\|2020-08-2 | ···················· | ···················· | ···················· | ···················· | ···················· | ···················· |
| Y | 7387 hCoV-19/Uganda/UG073/2020\|EPI_ISL_737942\|2020-08-2 | ···················· | ···················· | ···················· | ···················· | ···················· | ···················· |
| Y | 7388 hCoV-19/Uganda/UG074/2020\|EPI_ISL_737943\|2020-08-2 | ···················· | ···················· | ···················· | ···················· | ···················· | ···················· |
| Y | 7389 hCoV-19/Uganda/UG075/2020\|EPI_ISL_737944\|2020-08-2 | ···················· | ···················· | ···················· | ···················· | ···················· | ···················· |
| Y | 7390 hCoV-19/Uganda/UG100/2020\|EPI_ISL_737945\|2020-08-1 | ···················· | ···················· | ···················· | ···················· | ···················· | ···················· |
| Y | 7391 hCoV-19/Uganda/UG021/2020\|EPI_ISL_737946\|2020-05-0 | ···················· | ···················· | ···················· | ···················· | ···················· | ···················· |
| Y | 7392 hCoV-19/Uganda/UG022/2020\|EPI_ISL_737947\|2020-05-0 | ···················· | ···················· | ···················· | ···················· | ···················· | ···················· |
| Y | 7393 hCoV-19/Uganda/UG024/2020\|EPI_ISL_737948\|2020-05-0 | ···················· | ···················· | ···················· | ···················· | ···················· | ···················· |
| Y | 7394 hCoV-19/Uganda/UG025/2020\|EPI_ISL_737949\|2020-05-0 | ···················· | ···················· | ···················· | ···················· | ···················· | ···················· |
| Y | 7395 hCoV-19/Uganda/UG026/2020\|EPI_ISL_737950\|2020-05-0 | ···················· | ···················· | ···················· | ···················· | ···················· | ···················· |
| Y | 7396 hCoV-19/Uganda/UG027/2020\|EPI_ISL_737951\|2020-04-0 | ···················· | ···················· | ···················· | ···················· | ···················· | ···················· |
| Y | 7397 hCoV-19/Uganda/UG031/2020\|EPI_ISL_737952\|2020-05-0 | ···················· | ···················· | ···················· | ···················· | ···················· | ···················· |
| Y | 7398 hCoV-19/Uganda/UG033/2020\|EPI_ISL_737953\|2020-05-0 | ···················· | ···················· | ···················· | ···················· | ···················· | ···················· |
| Y | 7399 hCoV-19/Uganda/UG034/2020\|EPI_ISL_737954\|2020-03-2 | ···················· | ···················· | ···················· | ···················· | ···················· | ···················· |
| Y | 7400 hCoV-19/Uganda/UG035/2020\|EPI_ISL_737955\|2020-03-2 | ···················· | ···················· | ···················· | ···················· | ···················· | ···················· |
| Y | 7401 hCoV-19/Uganda/UG036/2020\|EPI_ISL_737956\|2020-03-3 | ···················· | ···················· | ···················· | ···················· | ···················· | ···················· |
| Y | 7402 hCoV-19/Uganda/UG037/2020\|EPI_ISL_737957\|2020-03-2 | ···················· | ···················· | ···················· | ···················· | ···················· | ···················· |
| Y | 7403 hCoV-19/Uganda/UG038/2020\|EPI_ISL_737958\|2020-03-2 | ···················· | ···················· | ···················· | ···················· | ···················· | ···················· |
| Y | 7404 hCoV-19/Uganda/UG039/2020\|EPI_ISL_737959\|2020-03-2 | ···················· | ···················· | ···················· | ···················· | ···················· | ···················· |
| Y | 7405 hCoV-19/Uganda/UG040/2020\|EPI_ISL_737960\|2020-05-0 | ···················· | ···················· | ···················· | ···················· | ···················· | ···················· |
| Y | 7406 hCoV-19/Uganda/UG041/2020\|EPI_ISL_737961\|2020-05-0 | ···················· | ···················· | ···················· | ···················· | ···················· | ···················· |
| Y | 7407 hCoV-19/Uganda/UG042/2020\|EPI_ISL_737962\|2020-05-0 | ···················· | ···················· | ···················· | ···················· | ···················· | ···················· |
| Y | 7408 hCoV-19/Uganda/UG043/2020\|EPI_ISL_737963\|2020-05-0 | ···················· | ···················· | ···················· | ···················· | ···················· | ···················· |
| Y | 7409 hCoV-19/Uganda/UG044/2020\|EPI_ISL_737964\|2020-05-0 | ···················· | ···················· | ···················· | ···················· | ···················· | ···················· |
| Y | 7410 hCoV-19/Uganda/UG045/2020\|EPI_ISL_737965\|2020-07-2 | ···················· | ···················· | ···················· | ···················· | ···················· | ···················· |
| Y | 7411 hCoV-19/Uganda/UG048/2020\|EPI_ISL_737966\|2020-05-0 | ···················· | ···················· | ···················· | ···················· | ···················· | ···················· |
| Y | 7412 hCoV-19/Uganda/UG049/2020\|EPI_ISL_737967\|2020-05-0 | ···················· | ···················· | ···················· | ···················· | ···················· | ···················· |
| Y | 7413 hCoV-19/Uganda/UG051/2020\|EPI_ISL_737968\|2020-05-0 | ···················· | ···················· | ···················· | ···················· | ···················· | ···················· |

| | | | | | | | | |
|---|---|---|---|---|---|---|---|---|
| Y | 7414 | hCoV-19/Uganda/UG052/2020\|EPI_ISL_737969\|2020-05-0 | ···················· | ···················· | ···················· | ···················· | ···················· | ···················· |
| Y | 7415 | hCoV-19/Uganda/UG053/2020\|EPI_ISL_737970\|2020-05-1 | ···················· | ···················· | ···················· | ···················· | ···················· | ···················· |
| Y | 7416 | hCoV-19/Uganda/UG054/2020\|EPI_ISL_737971\|2020-08-0 | ···················· | ···················· | ···················· | ···················· | ···················· | ···················· |
| Y | 7417 | hCoV-19/Uganda/UG055/2020\|EPI_ISL_737972\|2020-07-2 | ···················· | ···················· | ···················· | ···················· | ···················· | ···················· |
| Y | 7418 | hCoV-19/Uganda/UG056/2020\|EPI_ISL_737973\|2020-07-1 | ···················· | ···················· | ···················· | ···················· | ···················· | ···················· |
| Y | 7419 | hCoV-19/Uganda/UG057/2020\|EPI_ISL_737974\|2020-07-2 | ···················· | ···················· | ···················· | ···················· | ···················· | ···················· |
| Y | 7420 | hCoV-19/Uganda/UG059/2020\|EPI_ISL_737975\|2020-07-2 | ···················· | ···················· | ···················· | ···················· | ···················· | ···················· |
| Y | 7421 | hCoV-19/Uganda/UG060/2020\|EPI_ISL_737976\|2020-07-2 | ···················· | ···················· | ···················· | ···················· | ···················· | ···················· |
| Y | 7422 | hCoV-19/Uganda/UG061/2020\|EPI_ISL_737977\|2020-07-2 | ···················· | ·A·················· | ···················· | ···················· | ···················· | ···················· |
| Y | 7423 | hCoV-19/Uganda/UG062/2020\|EPI_ISL_737978\|2020-07-2 | ···················· | ···················· | ···················· | ···················· | ···················· | ···················· |
| Y | 7424 | hCoV-19/Uganda/UG063/2020\|EPI_ISL_737979\|2020-07-2 | ···················· | ···················· | ···················· | ···················· | ···················· | ···················· |
| Y | 7425 | hCoV-19/England/LOND-DAA01/2020\|EPI_ISL_724681\|202 | ···················· | ···················· | ···················· | ···················· | ···················· | ···················· |
| Y | 7426 | hCoV-19/England/LOND-DAA2F/2020\|EPI_ISL_724683\|202 | ···················· | ···················· | ···················· | ···················· | ···················· | ···················· |
| Y | 7427 | hCoV-19/England/LOND-DAA7A/2020\|EPI_ISL_724686\|202 | ···················· | ···················· | ···················· | ···················· | ···················· | ···················· |
| Y | 7428 | hCoV-19/England/LOND-DAA89/2020\|EPI_ISL_724687\|202 | ···················· | ···················· | ···················· | ···················· | ·····T·············· | ···················· |
| Y | 7429 | hCoV-19/Malaysia/IMR-WC258707/2020\|EPI_ISL_728210\| | ···················· | ·A·················· | ···················· | ···················· | ···················· | ···················· |
| Y | 7430 | hCoV-19/Malaysia/IMR-WC259750/2020\|EPI_ISL_728211\| | ···················· | ·A·················· | ···················· | ···················· | ···················· | ···················· |
| Y | 7431 | hCoV-19/Malaysia/IMR-WC268317/2020\|EPI_ISL_728212\| | ···················· | ·A·················· | ···················· | ···················· | ···················· | ···················· |
| Y | 7432 | hCoV-19/Morocco/LDB-02/2020\|EPI_ISL_728219\|2020-11 | ···················· | ···················· | ···················· | ···················· | ···················· | ···················· |
| Y | 7433 | hCoV-19/Germany/DE-HB-ChVir2097/2020\|EPI_ISL_72950 | ···················· | ···················· | ···················· | ···················· | ···················· | ···················· |
| Y | 7434 | hCoV-19/Germany/DE-NI-ChVir2098/2020\|EPI_ISL_72950 | ···················· | ···················· | ···················· | ···················· | ···················· | ···················· |
| Y | 7435 | hCoV-19/Germany/DE-NI-ChVir2099/2020\|EPI_ISL_72950 | ···················· | ···················· | ···················· | ···················· | ···················· | ···················· |
| Y | 7436 | hCoV-19/Germany/DE-HB-ChVir2101/2020\|EPI_ISL_72950 | ···················· | ···················· | ···················· | ···················· | ···················· | ···················· |
| Y | 7437 | hCoV-19/Germany/DE-SH-ChVir2105/2020\|EPI_ISL_72950 | ···················· | ···················· | ···················· | ···················· | ···················· | ···················· |
| Y | 7438 | hCoV-19/Germany/DE-SH-ChVir2109/2020\|EPI_ISL_72950 | ···················· | ···················· | ···················· | ···················· | ···················· | ···················· |
| Y | 7439 | hCoV-19/Germany/DE-SH-ChVir2111/2020\|EPI_ISL_72950 | ···················· | ···················· | ···················· | ···················· | ···················· | ···················· |
| Y | 7440 | hCoV-19/Germany/DE-SH-ChVir2114/2020\|EPI_ISL_72950 | ···················· | ···················· | ···················· | ···················· | ···················· | ···················· |
| Y | 7441 | hCoV-19/Germany/DE-SH-ChVir2117/2020\|EPI_ISL_72950 | ···················· | ···················· | ···················· | ···················· | ···················· | ···················· |
| Y | 7442 | hCoV-19/Germany/DE-SH-ChVir2118/2020\|EPI_ISL_72950 | ···················· | ···················· | ···················· | ···················· | ···················· | ···················· |
| Y | 7443 | hCoV-19/Germany/DE-SH-ChVir2119/2020\|EPI_ISL_72951 | ···················· | ···················· | ···················· | ···················· | ···················· | ···················· |
| Y | 7444 | hCoV-19/Germany/DE-SH-ChVir2123/2020\|EPI_ISL_72951 | ···················· | ···················· | ···················· | ···················· | ···················· | ···················· |
| Y | 7445 | hCoV-19/Germany/DE-SH-ChVir2125/2020\|EPI_ISL_72951 | ···················· | ···················· | ···················· | ···················· | ···················· | ···················· |
| Y | 7446 | hCoV-19/Germany/ChVir2127/2020\|EPI_ISL_729513\|2020 | ···················· | ···················· | ···················· | ···················· | ···················· | ···················· |
| Y | 7447 | hCoV-19/Germany/ChVir3001/2020\|EPI_ISL_729514\|2020 | ···················· | ···················· | ···················· | ···················· | ···················· | ···················· |
| Y | 7448 | hCoV-19/Germany/DE-SH-ChVir3032/2020\|EPI_ISL_72951 | ···················· | ···················· | ···················· | ···················· | ···················· | ···················· |
| Y | 7449 | hCoV-19/Germany/DE-SH-ChVir3034/2020\|EPI_ISL_72951 | ···················· | ···················· | ···················· | ···················· | ···················· | ···················· |
| Y | 7450 | hCoV-19/Germany/DE-SH-ChVir3035/2020\|EPI_ISL_72951 | ···················· | ···················· | ···················· | ···················· | ···················· | ···················· |
| Y | 7451 | hCoV-19/Germany/DE-SH-ChVir3037/2020\|EPI_ISL_72951 | ···················· | ···················· | ···················· | ···················· | ···················· | ···················· |
| Y | 7452 | hCoV-19/Germany/DE-SH-ChVir3038/2020\|EPI_ISL_72951 | ···················· | ···················· | ···················· | ···················· | ···················· | ···················· |
| Y | 7453 | hCoV-19/Germany/DE-SH-ChVir3196/2020\|EPI_ISL_72954 | ···················· | ···················· | ···················· | ···················· | ···················· | ···················· |
| Y | 7454 | hCoV-19/Germany/DE-SH-ChVir3200/2020\|EPI_ISL_72954 | ···················· | ···················· | ···················· | ···················· | ···················· | ···················· |
| Y | 7455 | hCoV-19/Germany/DE-SH-ChVir3201/2020\|EPI_ISL_72954 | ···················· | ···················· | ···················· | ···················· | ···················· | ···················· |
| Y | 7456 | hCoV-19/Germany/DE-SH-ChVir3203/2020\|EPI_ISL_72954 | ···················· | ···················· | ···················· | ···················· | ···················· | ···················· |
| Y | 7457 | hCoV-19/Germany/DE-SH-ChVir3214/2020\|EPI_ISL_72954 | ···················· | ···················· | ···················· | ···················· | ···················· | ···················· |
| Y | 7458 | hCoV-19/Germany/DE-SH-ChVir3215/2020\|EPI_ISL_72954 | ···················· | ···················· | ···················· | ···················· | ···················· | ···················· |
| Y | 7459 | hCoV-19/Germany/DE-SH-ChVir4733/2020\|EPI_ISL_72955 | ···················· | ···················· | ···················· | ···················· | ···················· | ···················· |
| Y | 7460 | hCoV-19/Germany/DE-SH-ChVir4734/2020\|EPI_ISL_72955 | ···················· | ···················· | ···················· | ···················· | ···················· | ···················· |
| Y | 7461 | hCoV-19/Germany/DE-SH-ChVir4764/2020\|EPI_ISL_72955 | ···················· | ···················· | ···················· | ···················· | ···················· | ···················· |
| Y | 7462 | hCoV-19/Germany/ChVir2009/2020\|EPI_ISL_729553\|2020 | ···················· | ···················· | ···················· | ···················· | ···················· | ···················· |
| Y | 7463 | hCoV-19/Germany/ChVir2012/2020\|EPI_ISL_729554\|2020 | ···················· | ···················· | ···················· | ···················· | ···················· | ···················· |
| Y | 7464 | hCoV-19/Germany/DE-BE-ChVir7621/2020\|EPI_ISL_72955 | ···················· | ···················· | ···················· | ···················· | ···················· | ···················· |
| Y | 7465 | hCoV-19/Germany/DE-BE-ChVir7622/2020\|EPI_ISL_72955 | ···················· | ···················· | ···················· | ···················· | ···················· | ···················· |
| Y | 7466 | hCoV-19/Germany/DE-BE-ChVir7623/2020\|EPI_ISL_72955 | ···················· | ···················· | ···················· | ···················· | ···················· | ···················· |
| Y | 7467 | hCoV-19/Germany/DE-BE-ChVir4227/2020\|EPI_ISL_72955 | ···················· | ···················· | ···················· | ···················· | ···················· | ···················· |
| Y | 7468 | hCoV-19/Germany/ChVir3423/2020\|EPI_ISL_729559\|2020 | ···················· | ···················· | ···················· | ···················· | ···················· | ···················· |
| Y | 7469 | hCoV-19/Jordan/AQ-ALSR-5262/2020\|EPI_ISL_730500\|20 | ···················· | ···················· | ···················· | ···················· | ···················· | ···················· |
| Y | 7470 | hCoV-19/Jordan/AM-ALSR-5286/2020\|EPI_ISL_730523\|20 | ···················· | ·A·················· | ···················· | ···················· | ···················· | ··A················· |
| Y | 7471 | hCoV-19/Jordan/AM-ALSR-5287/2020\|EPI_ISL_730524\|20 | ···················· | ·A·················· | ···················· | ···················· | ···················· | ··A················· |
| Y | 7472 | hCoV-19/Jordan/AM-ALSR-5288/2020\|EPI_ISL_730525\|20 | ···················· | ···················· | ···················· | ···················· | ···················· | ···················· |
| Y | 7473 | hCoV-19/Jordan/AM-ALSR-5289/2020\|EPI_ISL_730526\|20 | ···················· | ···················· | ···················· | ···················· | ···················· | ···················· |
| Y | 7474 | hCoV-19/Jordan/AM-ALSR-5290/2020\|EPI_ISL_730527\|20 | ···················· | ·A·················· | ···················· | ···················· | ···················· | ··A················· |
| Y | 7475 | hCoV-19/Jordan/AM-ALSR-5291/2020\|EPI_ISL_730528\|20 | ···················· | ···················· | ···················· | ···················· | ···················· | ···················· |
| Y | 7476 | hCoV-19/Jordan/AM-ALSR-5292/2020\|EPI_ISL_730529\|20 | ···················· | ·A·················· | ···················· | ···················· | ···················· | ··A················· |
| Y | 7477 | hCoV-19/Jordan/AM-ALSR-5293/2020\|EPI_ISL_730530\|20 | ···················· | ···················· | ···················· | ···················· | ···················· | ···················· |
| Y | 7478 | hCoV-19/Jordan/AM-ALSR-5296/2020\|EPI_ISL_730532\|20 | ···················· | ···················· | ···················· | ···················· | ···················· | ···················· |
| Y | 7479 | hCoV-19/Jordan/AZ-ALSR-5297/2020\|EPI_ISL_730533\|20 | ···················· | ·A·················· | ···················· | ···················· | ···················· | ··A················· |
| Y | 7480 | hCoV-19/Jordan/AM-ALSR-5298/2020\|EPI_ISL_730534\|20 | ···················· | ···················· | ···················· | ···················· | ···················· | ···················· |
| Y | 7481 | hCoV-19/Jordan/AM-ALSR-5299/2020\|EPI_ISL_730535\|20 | ···················· | ···················· | ···················· | ···················· | ···················· | ···················· |
| Y | 7482 | hCoV-19/Jordan/AM-ALSR-5300/2020\|EPI_ISL_730536\|20 | ···················· | ···················· | ···················· | ···················· | ···················· | ···················· |
| Y | 7483 | hCoV-19/Jordan/AZ-ALSR-5301/2020\|EPI_ISL_730537\|20 | ···················· | ···················· | ···················· | ···················· | ···················· | ···················· |
| Y | 7484 | hCoV-19/Jordan/IR-ALSR-5302/2020\|EPI_ISL_730538\|20 | ···················· | ···················· | ···················· | ···················· | ···················· | ···················· |
| Y | 7485 | hCoV-19/Jordan/AM-ALSR-5303/2020\|EPI_ISL_730539\|20 | ···················· | ···················· | ···················· | ···················· | ···················· | ···················· |
| Y | 7486 | hCoV-19/Jordan/AM-ALSR-5304/2020\|EPI_ISL_730540\|20 | ···················· | ···················· | ···················· | ···················· | ···················· | ···················· |
| Y | 7487 | hCoV-19/Jordan/AM-ALSR-5305/2020\|EPI_ISL_730541\|20 | ···················· | ···················· | ···················· | ···················· | ···················· | ···················· |
| Y | 7488 | hCoV-19/Jordan/AM-ALSR-5306/2020\|EPI_ISL_730542\|20 | ···················· | ···················· | ···················· | ···················· | ···················· | ···················· |
| Y | 7489 | hCoV-19/Jordan/AM-ALSR-5308/2020\|EPI_ISL_730544\|20 | ···················· | ·A·················· | ···················· | ···················· | ···················· | ··A················· |
| Y | 7490 | hCoV-19/England/ALDP-C3D44E/2020\|EPI_ISL_731801\|20 | ···················· | ···················· | ···················· | ···················· | ···················· | ···················· |
| Y | 7491 | hCoV-19/England/ALDP-C3B871/2020\|EPI_ISL_731802\|20 | ···················· | ···················· | ···················· | ···················· | ···················· | ···················· |
| Y | 7492 | hCoV-19/England/ALDP-C3D0FF/2020\|EPI_ISL_731803\|20 | ···················· | ···················· | ···················· | ···················· | ···················· | ···················· |
| Y | 7493 | hCoV-19/England/ALDP-C3CA6B/2020\|EPI_ISL_731804\|20 | ···················· | ···················· | ···················· | ···················· | ···················· | ···················· |

| | | | | | | | |
|---|---|---|---|---|---|---|---|
| Y | 7494 hCoV-19/England/ALDP-C3CAB6/2020\|EPI_ISL_731805\|20 | ···················· | ···················· | ···················· | ···················· | ···················· | ···················· |
| Y | 7495 hCoV-19/England/ALDP-C3B0A6/2020\|EPI_ISL_731806\|20 | ···················· | ···················· | ···················· | ···················· | ···················· | ···················· |
| Y | 7496 hCoV-19/England/ALDP-C3CBB3/2020\|EPI_ISL_731809\|20 | ···················· | ···················· | ···················· | ···················· | ···················· | ···················· |
| Y | 7497 hCoV-19/England/ALDP-C3B03D/2020\|EPI_ISL_731810\|20 | ···················· | ···················· | ···················· | ···················· | ···················· | ···················· |
| Y | 7498 hCoV-19/England/ALDP-C3DA88/2020\|EPI_ISL_731811\|20 | ···················· | ···················· | ···················· | ···················· | ···················· | ···················· |
| Y | 7499 hCoV-19/England/ALDP-C3D7BE/2020\|EPI_ISL_731812\|20 | ···················· | ···················· | ···················· | ···················· | ···················· | ···················· |
| Y | 7500 hCoV-19/England/ALDP-C3D99A/2020\|EPI_ISL_731813\|20 | ···················· | ···················· | ···················· | ···················· | ···················· | ···················· |
| Y | 7501 hCoV-19/England/ALDP-C3DDF8/2020\|EPI_ISL_731814\|20 | ···················· | ···················· | ···················· | ···················· | ···················· | ···················· |
| Y | 7502 hCoV-19/England/ALDP-C3CB2C/2020\|EPI_ISL_731815\|20 | ···················· | ···················· | ···················· | ···················· | ···················· | ···················· |
| Y | 7503 hCoV-19/England/ALDP-C3BE42/2020\|EPI_ISL_731816\|20 | ···················· | ···················· | ···················· | ···················· | ···················· | ···················· |
| Y | 7504 hCoV-19/England/ALDP-C3D36F/2020\|EPI_ISL_731817\|20 | ···················· | ···················· | ···················· | ···················· | ···················· | ···················· |
| Y | 7505 hCoV-19/England/ALDP-C3DAC4/2020\|EPI_ISL_731818\|20 | ···················· | ···················· | ···················· | ···················· | ···················· | ···················· |
| Y | 7506 hCoV-19/England/ALDP-C3C6A3/2020\|EPI_ISL_731820\|20 | ···················· | ···················· | ···················· | ···················· | ···················· | ···················· |
| Y | 7507 hCoV-19/England/ALDP-C3BBD2/2020\|EPI_ISL_731821\|20 | ···················· | ···················· | ···················· | ···················· | ···················· | ···················· |
| Y | 7508 hCoV-19/England/ALDP-C3D50F/2020\|EPI_ISL_731823\|20 | ···················· | ···················· | ···················· | ···················· | ···················· | ···················· |
| Y | 7509 hCoV-19/England/ALDP-C3DA97/2020\|EPI_ISL_731825\|20 | ···················· | ···················· | ···················· | ···················· | ···················· | ···················· |
| Y | 7510 hCoV-19/England/ALDP-C3D2AE/2020\|EPI_ISL_731826\|20 | ···················· | ···················· | ···················· | ···················· | ············R······· | ···················· |
| Y | 7511 hCoV-19/England/ALDP-C3B2A0/2020\|EPI_ISL_731827\|20 | ···················· | ···················· | ···················· | ···················· | ···················· | ···················· |
| Y | 7512 hCoV-19/England/ALDP-C3CBE0/2020\|EPI_ISL_731828\|20 | ···················· | ···················· | ···················· | ···················· | ···················· | ···················· |
| Y | 7513 hCoV-19/England/ALDP-C3D754/2020\|EPI_ISL_731829\|20 | ···················· | ···················· | ···················· | ···················· | ···················· | ···················· |
| Y | 7514 hCoV-19/England/ALDP-C3C685/2020\|EPI_ISL_731856\|20 | ···················· | ···················· | ···················· | ···················· | ···················· | ···················· |
| Y | 7515 hCoV-19/England/ALDP-C3ABE2/2020\|EPI_ISL_731858\|20 | ···················· | ···················· | ···················· | ···················· | ···················· | ···················· |
| Y | 7516 hCoV-19/England/ALDP-C3C8AD/2020\|EPI_ISL_731859\|20 | ···················· | ···················· | ···················· | ···················· | ···················· | ···················· |
| Y | 7517 hCoV-19/England/ALDP-C3CE50/2020\|EPI_ISL_731860\|20 | ···················· | ···················· | ···················· | ···················· | ···················· | ···················· |
| Y | 7518 hCoV-19/England/ALDP-C3AC0D/2020\|EPI_ISL_731863\|20 | ···················· | ···················· | ···················· | ···················· | ···················· | ···················· |
| Y | 7519 hCoV-19/England/ALDP-C3BE24/2020\|EPI_ISL_731864\|20 | ···················· | ···················· | ···················· | ···················· | ···················· | ···················· |
| Y | 7520 hCoV-19/England/ALDP-C3D43F/2020\|EPI_ISL_731867\|20 | ···················· | ···················· | ···················· | ···················· | ···················· | ···················· |
| Y | 7521 hCoV-19/England/ALDP-C3B49B/2020\|EPI_ISL_731868\|20 | ···················· | ···················· | ···················· | ···················· | ···················· | ···················· |
| Y | 7522 hCoV-19/England/ALDP-C3DD34/2020\|EPI_ISL_731869\|20 | ···················· | ···················· | ···················· | ···················· | ······T············· | ···················· |
| Y | 7523 hCoV-19/England/ALDP-C3D341/2020\|EPI_ISL_731870\|20 | ···················· | ···················· | ···················· | ···················· | ···················· | ···················· |
| Y | 7524 hCoV-19/England/ALDP-C3CD9F/2020\|EPI_ISL_731871\|20 | ···················· | ···················· | ···················· | ···················· | ···················· | ···················· |
| Y | 7525 hCoV-19/England/ALDP-C3BB69/2020\|EPI_ISL_731873\|20 | ···················· | ···················· | ···················· | ···················· | ···················· | ···················· |
| Y | 7526 hCoV-19/England/ALDP-C3AAA9/2020\|EPI_ISL_731874\|20 | ···················· | ···················· | ···················· | ···················· | ···················· | ···················· |
| Y | 7527 hCoV-19/England/ALDP-C3B158/2020\|EPI_ISL_731875\|20 | ···················· | ···················· | ···················· | ···················· | ···················· | ···················· |
| Y | 7528 hCoV-19/England/QEUH-C32E1E/2020\|EPI_ISL_734000\|20 | ···················· | ···················· | ···················· | ···················· | ······T············· | ···················· |
| Y | 7529 hCoV-19/England/QEUH-C33A37/2020\|EPI_ISL_734001\|20 | ···················· | ···················· | ···················· | ···················· | ···················· | ···················· |
| Y | 7530 hCoV-19/England/QEUH-C33EFF/2020\|EPI_ISL_734002\|20 | ···················· | ···················· | ···················· | ···················· | ···················· | ···················· |
| Y | 7531 hCoV-19/England/QEUH-C333D2/2020\|EPI_ISL_734005\|20 | ···················· | ···················· | ···················· | ···················· | ···················· | ···················· |
| Y | 7532 hCoV-19/England/QEUH-C33141/2020\|EPI_ISL_734006\|20 | ···················· | ···················· | ···················· | ···················· | ···················· | ···················· |
| Y | 7533 hCoV-19/England/QEUH-C33B70/2020\|EPI_ISL_734009\|20 | ···················· | ···················· | ···················· | ···················· | ···················· | ···················· |
| Y | 7534 hCoV-19/England/QEUH-C34483/2020\|EPI_ISL_734011\|20 | ···················· | ···················· | ···················· | ···················· | ···················· | ···················· |
| Y | 7535 hCoV-19/England/QEUH-C33E2C/2020\|EPI_ISL_734012\|20 | ···················· | ···················· | ···················· | ···················· | ···················· | ···················· |
| Y | 7536 hCoV-19/England/QEUH-C3440B/2020\|EPI_ISL_734014\|20 | ···················· | ···················· | ···················· | ···················· | ···················· | ···················· |
| Y | 7537 hCoV-19/England/QEUH-C3469C/2020\|EPI_ISL_734015\|20 | ···················· | ···················· | ···················· | ···················· | ···················· | ···················· |
| Y | 7538 hCoV-19/England/QEUH-C33B16/2020\|EPI_ISL_734031\|20 | ···················· | ···················· | ···················· | ···················· | ···················· | ···················· |
| Y | 7539 hCoV-19/England/QEUH-C34043/2020\|EPI_ISL_734032\|20 | ···················· | ···················· | ···················· | ···················· | ···················· | ···················· |
| Y | 7540 hCoV-19/England/QEUH-C32ED2/2020\|EPI_ISL_734033\|20 | ···················· | ···················· | ···················· | ···················· | ···················· | ···················· |
| Y | 7541 hCoV-19/England/QEUH-C3480F/2020\|EPI_ISL_734034\|20 | ···················· | ···················· | ···················· | ···················· | ···················· | ···················· |
| Y | 7542 hCoV-19/England/QEUH-C32F57/2020\|EPI_ISL_734035\|20 | ···················· | ···················· | ···················· | ···················· | ···················· | ···················· |
| Y | 7543 hCoV-19/England/QEUH-C32F48/2020\|EPI_ISL_734036\|20 | ···················· | ···················· | ···················· | ···················· | ···················· | ···················· |
| Y | 7544 hCoV-19/England/QEUH-C3390D/2020\|EPI_ISL_734037\|20 | ···················· | ···················· | ···················· | ···················· | ···················· | ···A················ |
| Y | 7545 hCoV-19/England/QEUH-C34104/2020\|EPI_ISL_734038\|20 | ···················· | ···················· | ···················· | ···················· | ···················· | ···················· |
| Y | 7546 hCoV-19/England/QEUH-C34614/2020\|EPI_ISL_734039\|20 | ···················· | ···················· | ···················· | ···················· | ···················· | ···················· |
| Y | 7547 hCoV-19/England/QEUH-C331C9/2020\|EPI_ISL_734040\|20 | ···················· | ···················· | ···················· | ···················· | ···················· | ···················· |
| Y | 7548 hCoV-19/England/QEUH-C3424D/2020\|EPI_ISL_734044\|20 | ···················· | ···················· | ···················· | ···················· | ···················· | ···················· |
| Y | 7549 hCoV-19/England/QEUH-C337B8/2020\|EPI_ISL_734045\|20 | ···················· | ···················· | ···················· | ···················· | ···················· | ···················· |
| Y | 7550 hCoV-19/England/QEUH-C33378/2020\|EPI_ISL_734046\|20 | ···················· | ···················· | ···················· | ···················· | ···················· | ···················· |
| Y | 7551 hCoV-19/England/QEUH-C345BD/2020\|EPI_ISL_734047\|20 | ···················· | ···················· | ···················· | ···················· | ···················· | ···················· |
| Y | 7552 hCoV-19/England/QEUH-C33D01/2020\|EPI_ISL_734050\|20 | ···················· | ···················· | ···················· | ···················· | ···················· | ···················· |
| Y | 7553 hCoV-19/England/QEUH-C3335A/2020\|EPI_ISL_734052\|20 | ···················· | ···················· | ···················· | ···················· | ···················· | ···················· |
| Y | 7554 hCoV-19/England/QEUH-C3478A/2020\|EPI_ISL_734053\|20 | ···················· | ···················· | ···················· | ···················· | ···················· | ···················· |
| Y | 7555 hCoV-19/England/QEUH-C346AB/2020\|EPI_ISL_734054\|20 | ···················· | ···················· | ···················· | ···················· | ···················· | ···················· |
| Y | 7556 hCoV-19/England/QEUH-C33EC2/2020\|EPI_ISL_734056\|20 | ···················· | ···················· | ···················· | ···················· | ···················· | ···················· |
| Y | 7557 hCoV-19/England/QEUH-C34702/2020\|EPI_ISL_734057\|20 | ···················· | ···················· | ···················· | ···················· | ···················· | ···················· |
| Y | 7558 hCoV-19/England/QEUH-C34623/2020\|EPI_ISL_734058\|20 | ···················· | ···················· | ···················· | ···················· | ······T············· | ···················· |
| Y | 7559 hCoV-19/England/QEUH-C347A8/2020\|EPI_ISL_734059\|20 | ···················· | ···················· | ···················· | ···················· | ···················· | ···················· |
| Y | 7560 hCoV-19/England/QEUH-C33D10/2020\|EPI_ISL_734060\|20 | ···················· | ···················· | ···················· | ···················· | ···················· | ···················· |
| Y | 7561 hCoV-19/England/QEUH-C333C3/2020\|EPI_ISL_734061\|20 | ···················· | ·A·················· | ···················· | ···················· | ···················· | ···················· |
| Y | 7562 hCoV-19/England/QEUH-C33A91/2020\|EPI_ISL_734062\|20 | ···················· | ···················· | ···················· | ···················· | ···················· | ···················· |
| Y | 7563 hCoV-19/England/QEUH-C34456/2020\|EPI_ISL_734065\|20 | ···················· | ···················· | ···················· | ···················· | ···················· | ···················· |
| Y | 7564 hCoV-19/England/QEUH-C34896/2020\|EPI_ISL_734068\|20 | ···················· | ···················· | ···················· | ···················· | ···················· | ···················· |
| Y | 7565 hCoV-19/England/QEUH-C34289/2020\|EPI_ISL_734070\|20 | ···················· | ···················· | ···················· | ···················· | ···················· | ···················· |
| Y | 7566 hCoV-19/England/QEUH-C3409E/2020\|EPI_ISL_734071\|20 | ···················· | ···················· | ···················· | ···················· | ······T············· | ···················· |
| Y | 7567 hCoV-19/England/QEUH-C32F66/2020\|EPI_ISL_734072\|20 | ···················· | ···················· | ···················· | ···················· | ···················· | ···················· |
| Y | 7568 hCoV-19/England/QEUH-C33D98/2020\|EPI_ISL_734073\|20 | ···················· | ···················· | ···················· | ···················· | ······T············· | ···················· |
| Y | 7569 hCoV-19/England/QEUH-C33E59/2020\|EPI_ISL_734074\|20 | ···················· | ···················· | ···················· | ···················· | ···················· | ···················· |
| Y | 7570 hCoV-19/England/QEUH-C33132/2020\|EPI_ISL_734075\|20 | ···················· | ···················· | ···················· | ···················· | ···················· | ···················· |
| Y | 7571 hCoV-19/England/QEUH-C34052/2020\|EPI_ISL_734076\|20 | ···················· | ···················· | ···················· | ···················· | ···················· | ···················· |
| Y | 7572 hCoV-19/England/QEUH-C33387/2020\|EPI_ISL_734077\|20 | ···················· | ···················· | ···················· | ···················· | ···················· | ···················· |
| Y | 7573 hCoV-19/England/QEUH-C33B43/2020\|EPI_ISL_734078\|20 | ···················· | ···················· | ···················· | ···················· | ···················· | ···················· |

| | | | | | | | | |
|---|---|---|---|---|---|---|---|---|
| Y | 7574 | hCoV-19/England/QEUH-C33C9B/2020\|EPI_ISL_734079\|20 | ···················· | ···················· | ···················· | ···················· | ···················· | ···················· |
| Y | 7575 | hCoV-19/England/QEUH-C33C40/2020\|EPI_ISL_734080\|20 | ···················· | ···················· | ···················· | ···················· | ···················· | ···················· |
| Y | 7576 | hCoV-19/England/QEUH-C343C2/2020\|EPI_ISL_734081\|20 | ···················· | ···················· | ···················· | ···················· | ···················· | ···················· |
| Y | 7577 | hCoV-19/England/QEUH-C3331E/2020\|EPI_ISL_734083\|20 | ···················· | ·········T·········· | ···················· | ···················· | ···················· | ···················· |
| Y | 7578 | hCoV-19/England/QEUH-C3375E/2020\|EPI_ISL_734084\|20 | ···················· | ···················· | ···················· | ···················· | ···················· | ···················· |
| Y | 7579 | hCoV-19/England/QEUH-C3459F/2020\|EPI_ISL_734086\|20 | ···················· | ···················· | ···················· | ···················· | ···················· | ···················· |
| Y | 7580 | hCoV-19/England/QEUH-C32F84/2020\|EPI_ISL_734087\|20 | ···················· | ·A·················· | ···················· | ···················· | ···················· | ···················· |
| Y | 7581 | hCoV-19/England/QEUH-C3434A/2020\|EPI_ISL_734088\|20 | ···················· | ···················· | ···················· | ···················· | ···················· | ···················· |
| Y | 7582 | hCoV-19/England/QEUH-C32F75/2020\|EPI_ISL_734089\|20 | ···················· | ···················· | ···················· | ···················· | ···················· | ···················· |
| Y | 7583 | hCoV-19/England/QEUH-C33B25/2020\|EPI_ISL_734094\|20 | ···················· | ···················· | ···················· | ···················· | ···················· | ···················· |
| Y | 7584 | hCoV-19/England/QEUH-C34447/2020\|EPI_ISL_734095\|20 | ···················· | ···················· | ···················· | ···················· | ···················· | ···················· |
| Y | 7585 | hCoV-19/England/QEUH-C33949/2020\|EPI_ISL_734096\|20 | ···················· | ···················· | ···················· | ···················· | ···················· | ···················· |
| Y | 7586 | hCoV-19/England/QEUH-C34429/2020\|EPI_ISL_734097\|20 | ···················· | ···················· | ···················· | ···················· | ···················· | ···················· |
| Y | 7587 | hCoV-19/England/QEUH-C32E87/2020\|EPI_ISL_734098\|20 | ···················· | ···················· | ···················· | ···················· | ···················· | ···················· |
| Y | 7588 | hCoV-19/England/QEUH-C33FCF/2020\|EPI_ISL_734099\|20 | ···················· | ···················· | ···················· | ···················· | ···················· | ···················· |
| Y | 7589 | hCoV-19/Turkey/IST-GLAB-254/2020\|EPI_ISL_735319\|20 | ···················· | ···················· | ···················· | ···················· | ···················· | ···················· |
| Y | 7590 | hCoV-19/Turkey/IST-GLAB-255/2020\|EPI_ISL_735320\|20 | ···················· | ···················· | ···················· | ···················· | ···················· | ···················· |
| Y | 7591 | hCoV-19/Turkey/IST-GLAB-258/2020\|EPI_ISL_735321\|20 | ···················· | ···················· | ···················· | ···················· | ···················· | ···················· |
| Y | 7592 | hCoV-19/Turkey/IST-GLAB-261/2020\|EPI_ISL_735322\|20 | ···················· | ···················· | ···················· | ···················· | ···················· | ···················· |
| Y | 7593 | hCoV-19/Turkey/IST-GLAB-263/2020\|EPI_ISL_735323\|20 | ···················· | ···················· | ···················· | ···················· | ···················· | ···················· |
| Y | 7594 | hCoV-19/Turkey/IST-GLAB-265/2020\|EPI_ISL_735324\|20 | ···················· | ···················· | ···················· | ···················· | ···················· | ···················· |
| Y | 7595 | hCoV-19/Turkey/IST-GLAB-267/2020\|EPI_ISL_735325\|20 | ···················· | ···················· | ···················· | ···················· | ···················· | ···················· |
| Y | 7596 | hCoV-19/Turkey/IST-GLAB-269/2020\|EPI_ISL_735326\|20 | ···················· | ···················· | ···················· | ···················· | ···················· | ···················· |
| Y | 7597 | hCoV-19/Turkey/IST-GLAB-272/2020\|EPI_ISL_735327\|20 | ···················· | ···················· | ···················· | ···················· | ···················· | ···················· |
| Y | 7598 | hCoV-19/Turkey/IST-GLAB-273/2020\|EPI_ISL_735328\|20 | ···················· | ···················· | ···················· | ···················· | ···················· | ···················· |
| Y | 7599 | hCoV-19/Turkey/IST-GLAB-274/2020\|EPI_ISL_735329\|20 | ···················· | ···················· | ···················· | ···················· | ···················· | ···················· |
| Y | 7600 | hCoV-19/Turkey/IST-GLAB-282/2020\|EPI_ISL_735330\|20 | ···················· | ···················· | ···················· | ···················· | ···················· | ···················· |
| Y | 7601 | hCoV-19/Turkey/IST-GLAB-283/2020\|EPI_ISL_735331\|20 | ···················· | ···················· | ···················· | ···················· | ···················· | ···················· |
| Y | 7602 | hCoV-19/Turkey/IST-GLAB-284/2020\|EPI_ISL_735332\|20 | ···················· | ···················· | ···················· | ···················· | ···················· | ···················· |
| Y | 7603 | hCoV-19/Turkey/IST-GLAB-287/2020\|EPI_ISL_735333\|20 | ···················· | ···················· | ···················· | ···················· | ···················· | ···················· |
| Y | 7604 | hCoV-19/Turkey/IST-GLAB-289/2020\|EPI_ISL_735334\|20 | ···················· | ·A·················· | ···················· | ···················· | ···················· | ···A················ |
| Y | 7605 | hCoV-19/Turkey/IST-GLAB-290/2020\|EPI_ISL_735335\|20 | ···················· | ······A············· | ···················· | ···················· | ···················· | ···················· |
| Y | 7606 | hCoV-19/Turkey/IST-GLAB-291/2020\|EPI_ISL_735336\|20 | ···················· | ···················· | ···················· | ···················· | ···················· | ···················· |
| Y | 7607 | hCoV-19/Turkey/IST-GLAB-293/2020\|EPI_ISL_735337\|20 | ···················· | ···················· | ···················· | ···················· | ···················· | ···················· |
| Y | 7608 | hCoV-19/Turkey/IST-GLAB-295/2020\|EPI_ISL_735338\|20 | ···················· | ···················· | ···················· | ···················· | ···················· | ···················· |
| Y | 7609 | hCoV-19/Turkey/IST-GLAB-299/2020\|EPI_ISL_735339\|20 | ···················· | ···················· | ···················· | ···················· | ···················· | ···················· |
| Y | 7610 | hCoV-19/Turkey/IST-GLAB-300/2020\|EPI_ISL_735340\|20 | ···················· | ···················· | ···················· | ···················· | ···················· | ···················· |
| Y | 7611 | hCoV-19/Turkey/IST-GLAB-302/2020\|EPI_ISL_735342\|20 | ···················· | ···················· | ···················· | ···················· | ···················· | ···················· |
| Y | 7612 | hCoV-19/Turkey/IST-GLAB-303/2020\|EPI_ISL_735343\|20 | ···················· | ···················· | ···················· | ···················· | ···················· | ···················· |
| Y | 7613 | hCoV-19/Turkey/IST-GLAB-312/2020\|EPI_ISL_735346\|20 | ···················· | ···················· | ···················· | ···················· | ···················· | ···················· |
| Y | 7614 | hCoV-19/Turkey/IST-GLAB-313/2020\|EPI_ISL_735347\|20 | ···················· | ···················· | ···················· | ···················· | ···················· | ···················· |
| Y | 7615 | hCoV-19/Turkey/IST-GLAB-348/2020\|EPI_ISL_735360\|20 | ···················· | ·A·················· | ···················· | ···················· | ···················· | ···················· |
| Y | 7616 | hCoV-19/Turkey/IST-GLAB-349/2020\|EPI_ISL_735361\|20 | ···················· | ···················· | ···················· | ···················· | ···················· | ···················· |
| Y | 7617 | hCoV-19/Turkey/IST-GLAB-350/2020\|EPI_ISL_735362\|20 | ···················· | ···················· | ···················· | ···················· | ···················· | ···················· |
| Y | 7618 | hCoV-19/Turkey/IST-GLAB-351/2020\|EPI_ISL_735363\|20 | ···················· | ···················· | ···················· | ···················· | ···················· | ···················· |
| Y | 7619 | hCoV-19/Turkey/IST-GLAB-352/2020\|EPI_ISL_735364\|20 | ···················· | ···················· | ···················· | ···················· | ···················· | ···················· |
| Y | 7620 | hCoV-19/Turkey/IST-GLAB-353/2020\|EPI_ISL_735365\|20 | ···················· | ···················· | ···················· | ···················· | ···················· | ···················· |
| Y | 7621 | hCoV-19/Turkey/IST-GLAB-A1/2020\|EPI_ISL_735366\|202 | ···················· | ···················· | ···················· | ···················· | ···················· | ···················· |
| Y | 7622 | hCoV-19/Turkey/IST-GLAB-A10/2020\|EPI_ISL_735367\|20 | ···················· | ···················· | ···················· | ···················· | ···················· | ···················· |
| Y | 7623 | hCoV-19/Turkey/IST-GLAB-A3/2020\|EPI_ISL_735368\|202 | ···················· | ···················· | ···················· | ···················· | ···················· | ···················· |
| Y | 7624 | hCoV-19/Turkey/IST-GLAB-A4/2020\|EPI_ISL_735369\|202 | ···················· | ···················· | ···················· | ···················· | ···················· | ···················· |
| Y | 7625 | hCoV-19/Turkey/IST-GLAB-A5/2020\|EPI_ISL_735370\|202 | ···················· | ···················· | ···················· | ···················· | ···················· | ···················· |
| Y | 7626 | hCoV-19/Turkey/IST-GLAB-A6/2020\|EPI_ISL_735371\|202 | ···················· | ···················· | ···················· | ···················· | ···················· | ···················· |
| Y | 7627 | hCoV-19/Turkey/IST-GLAB-A9/2020\|EPI_ISL_735373\|202 | ···················· | ···················· | ···················· | ···················· | ···················· | ···················· |
| Y | 7628 | hCoV-19/USA/CA-LACPHL-AE00043/2020\|EPI_ISL_735374\| | ···················· | ····T··············· | ···················· | ···················· | ···················· | ···················· |
| Y | 7629 | hCoV-19/USA/CA-LACPHL-AE00044/2020\|EPI_ISL_735375\| | ···················· | ···················· | ···················· | ···················· | ···················· | ···················· |
| Y | 7630 | hCoV-19/USA/CA-LACPHL-AE00045/2020\|EPI_ISL_735376\| | ···················· | ····T··············· | ···················· | ···················· | ···················· | ···················· |
| Y | 7631 | hCoV-19/USA/CA-LACPHL-AE00046/2020\|EPI_ISL_735377\| | ···················· | ·A·················· | ···················· | ···················· | ···················· | ···················· |
| Y | 7632 | hCoV-19/USA/CA-LACPHL-AE00049/2020\|EPI_ISL_735378\| | ···················· | ···················· | ···················· | ···················· | ···················· | ···················· |
| Y | 7633 | hCoV-19/USA/CA-LACPHL-AE00050/2020\|EPI_ISL_735379\| | ···················· | ····T··············· | ···················· | ···················· | ···················· | ···················· |
| Y | 7634 | hCoV-19/USA/CA-LACPHL-AE00051/2020\|EPI_ISL_735380\| | ···················· | ···················· | ···················· | ···················· | ···················· | ···················· |
| Y | 7635 | hCoV-19/USA/CA-LACPHL-AE00052/2020\|EPI_ISL_735381\| | ···················· | ····T··············· | ···················· | ···················· | ···················· | ···················· |
| Y | 7636 | hCoV-19/USA/CA-LACPHL-AE00053/2020\|EPI_ISL_735382\| | ···················· | ····T··············· | ···················· | ···················· | ···················· | ···················· |
| Y | 7637 | hCoV-19/USA/CA-LACPHL-AE00054/2020\|EPI_ISL_735383\| | ···················· | ····T··············· | ···················· | ···················· | ···················· | ···················· |
| Y | 7638 | hCoV-19/Brazil/SP-585/2020\|EPI_ISL_735396\|2020-07- | ···················· | ···················· | ···················· | ···················· | ···················· | ···················· |
| Y | 7639 | hCoV-19/Brazil/SP-586/2020\|EPI_ISL_735397\|2020-07- | ···················· | ···················· | ···················· | ···················· | ···················· | ···················· |
| Y | 7640 | hCoV-19/Brazil/SP-737/2020\|EPI_ISL_735398\|2020-07- | ···················· | ···················· | ···················· | ···················· | ···················· | ···················· |
| Y | 7641 | hCoV-19/Brazil/SP-738/2020\|EPI_ISL_735399\|2020-04- | ···················· | ···················· | ···················· | ···················· | ···················· | ···················· |
| Y | 7642 | hCoV-19/England/QEUH-C4BD60/2020\|EPI_ISL_736650\|20 | ···················· | ···················· | ···················· | ···················· | ···················· | ···················· |
| Y | 7643 | hCoV-19/England/ALDP-C48CA2/2020\|EPI_ISL_736652\|20 | ···················· | ···················· | ···················· | ···················· | ···················· | ···················· |
| Y | 7644 | hCoV-19/England/ALDP-C4898D/2020\|EPI_ISL_736654\|20 | ···················· | ···················· | ···················· | ···················· | ···················· | ···················· |
| Y | 7645 | hCoV-19/England/ALDP-C48CEE/2020\|EPI_ISL_736655\|20 | ···················· | ···················· | ···················· | ···················· | ···················· | ···················· |
| Y | 7646 | hCoV-19/England/ALDP-C485F2/2020\|EPI_ISL_736656\|20 | ···················· | ···················· | ···················· | ···················· | ···················· | ···················· |
| Y | 7647 | hCoV-19/England/QEUH-C4BD42/2020\|EPI_ISL_736657\|20 | ···················· | ···················· | ···················· | ···················· | ···················· | ···················· |
| Y | 7648 | hCoV-19/England/ALDP-C48AA8/2020\|EPI_ISL_736661\|20 | ···················· | ···················· | ···················· | ···················· | ···················· | ···················· |
| Y | 7649 | hCoV-19/England/ALDP-C486FF/2020\|EPI_ISL_736662\|20 | ···················· | ·A·················· | ···················· | ···················· | ···················· | ···A················ |
| Y | 7650 | hCoV-19/England/ALDP-C4848C/2020\|EPI_ISL_736664\|20 | ···················· | ···················· | ···················· | ···················· | ···················· | ···················· |
| Y | 7651 | hCoV-19/England/ALDP-C48BD2/2020\|EPI_ISL_736665\|20 | ···················· | ···················· | ···················· | ···················· | ···················· | ···················· |
| Y | 7652 | hCoV-19/England/ALDP-C48D27/2020\|EPI_ISL_736666\|20 | ···················· | ···················· | ···················· | ···················· | ···················· | ···················· |
| Y | 7653 | hCoV-19/England/ALDP-C48CFD/2020\|EPI_ISL_736667\|20 | ···················· | ···················· | ···················· | ···················· | ···················· | ···················· |

| | | | | | | | |
|---|---|---|---|---|---|---|---|
| Y | 7654 hCoV-19/England/ALDP-C48DDC/2020\|EPI_ISL_736670\|20 | ···················· | ···················· | ····················· | ···················· | ···················· | ···················· |
| Y | 7655 hCoV-19/England/QEUH-C4BBFD/2020\|EPI_ISL_736672\|20 | ···················· | ···················· | ····················· | ···················· | ···················· | ···················· |
| Y | 7656 hCoV-19/England/MILK-C5059D/2020\|EPI_ISL_736675\|20 | ···················· | ···················· | ····················· | ···················· | ···················· | ···················· |
| Y | 7657 hCoV-19/England/QEUH-C4BBC0/2020\|EPI_ISL_736678\|20 | ···················· | ···················· | ····················· | ···················· | ···················· | ···················· |
| Y | 7658 hCoV-19/England/MILK-C50515/2020\|EPI_ISL_736693\|20 | ···················· | ···················· | ····················· | ···················· | ···················· | ···················· |
| Y | 7659 hCoV-19/England/QEUH-C4BD06/2020\|EPI_ISL_736695\|20 | ···················· | ···················· | ····················· | ···················· | ···················· | ···················· |
| Y | 7660 hCoV-19/England/ALDP-C48D09/2020\|EPI_ISL_736698\|20 | ···················· | ···················· | ····················· | ···················· | ···················· | ···················· |
| Y | 7661 hCoV-19/Uganda/UG064/2020\|EPI_ISL_737980\|2020-08-2 | ···················· | ···················· | ····················· | ···················· | ···················· | ···················· |
| Y | 7662 hCoV-19/Uganda/UG065/2020\|EPI_ISL_737981\|2020-08-2 | ···················· | ···················· | ····················· | ···················· | ···················· | ···················· |
| Y | 7663 hCoV-19/Uganda/UG067/2020\|EPI_ISL_737982\|2020-08-2 | ···················· | ···················· | ····················· | ···················· | ···················· | ···················· |
| Y | 7664 hCoV-19/Uganda/UG069/2020\|EPI_ISL_737983\|2020-08-2 | ···················· | ···················· | ····················· | ···················· | ···················· | ···················· |
| Y | 7665 hCoV-19/Uganda/UG070/2020\|EPI_ISL_737984\|2020 | ···················· | ···················· | ····················· | ···················· | ···················· | ···················· |
| Y | 7666 hCoV-19/Uganda/UG071/2020\|EPI_ISL_737985\|2020-08-2 | ···················· | ···················· | ····················· | ···················· | ···················· | ···················· |
| Y | 7667 hCoV-19/Uganda/UG072/2020\|EPI_ISL_737986\|2020-08-2 | ···················· | ···················· | ····················· | ···················· | ···················· | ···················· |
| Y | 7668 hCoV-19/Uganda/UG076/2020\|EPI_ISL_737987\|2020-08-2 | ···················· | ···················· | ····················· | ···················· | ···················· | ···················· |
| Y | 7669 hCoV-19/Uganda/UG077/2020\|EPI_ISL_737988\|2020-08-3 | ···················· | ···················· | ····················· | ···················· | ···················· | ···················· |
| Y | 7670 hCoV-19/Uganda/UG078/2020\|EPI_ISL_737989\|2020-08-2 | ···················· | ···················· | ····················· | ···················· | ···················· | ···················· |
| Y | 7671 hCoV-19/Uganda/UG079/2020\|EPI_ISL_737990\|2020-09-0 | ···················· | ···················· | ····················· | ···················· | ···················· | ···················· |
| Y | 7672 hCoV-19/Uganda/UG080/2020\|EPI_ISL_737991\|2020-08-1 | ···················· | ···················· | ····················· | ···················· | ···················· | ···················· |
| Y | 7673 hCoV-19/Uganda/UG081/2020\|EPI_ISL_737992\|2020-08-1 | ···················· | ···················· | ····················· | ···················· | ···················· | ···················· |
| Y | 7674 hCoV-19/Uganda/UG082/2020\|EPI_ISL_737993\|2020-08-1 | ···················· | ···················· | ····················· | ···················· | ···················· | ···················· |
| Y | 7675 hCoV-19/Uganda/UG083/2020\|EPI_ISL_737994\|2020-08-1 | ···················· | ···················· | ····················· | ···················· | ···················· | ···················· |
| Y | 7676 hCoV-19/Uganda/UG084/2020\|EPI_ISL_737995\|2020-08-1 | ···················· | ···················· | ····················· | ···················· | ···················· | ···················· |
| Y | 7677 hCoV-19/Uganda/UG085/2020\|EPI_ISL_737996\|2020 | ···················· | ···················· | ····················· | ···················· | ···················· | ···················· |
| Y | 7678 hCoV-19/Uganda/UG086/2020\|EPI_ISL_737997\|2020-08-2 | ···················· | ···················· | ····················· | ···················· | ···················· | ···················· |
| Y | 7679 hCoV-19/Uganda/UG087/2020\|EPI_ISL_737998\|2020-08-2 | ···················· | ···················· | ····················· | ···················· | ···················· | ···················· |
| Y | 7680 hCoV-19/Uganda/UG088/2020\|EPI_ISL_737999\|2020-08-2 | ···················· | ···················· | ····················· | ···················· | ···················· | ···················· |
| Y | 7681 hCoV-19/England/LOND-DADEA/2020\|EPI_ISL_724713\|202 | ···················· | ···················· | ····················· | ···················· | ···················· | ···················· |
| Y | 7682 hCoV-19/England/LOND-DAE8D/2020\|EPI_ISL_724722\|202 | ···················· | ···················· | ····················· | ···················· | ···················· | ···················· |
| Y | 7683 hCoV-19/England/LOND-DAE9C/2020\|EPI_ISL_724723\|202 | ···················· | ···················· | ····················· | ···················· | ···················· | ···················· |
| Y | 7684 hCoV-19/England/LOND-DAEBA/2020\|EPI_ISL_724724\|202 | ···················· | ···················· | ····················· | ···················· | ···················· | ···················· |
| Y | 7685 hCoV-19/England/LOND-DAEC9/2020\|EPI_ISL_724725\|202 | ···················· | ···················· | ····················· | ···················· | ···················· | ···················· |
| Y | 7686 hCoV-19/England/LOND-DAEF6/2020\|EPI_ISL_724728\|202 | ···················· | ···················· | ····················· | ···················· | ···················· | ···················· |
| Y | 7687 hCoV-19/England/LOND-DAF3F/2020\|EPI_ISL_724732\|202 | ···················· | ···················· | ····················· | ···················· | ···················· | ···················· |
| Y | 7688 hCoV-19/England/LOND-DB244/2020\|EPI_ISL_724747\|202 | ···················· | ···················· | ····················· | ···················· | ···················· | ···················· |
| Y | 7689 hCoV-19/England/LOND-DB4A8/2020\|EPI_ISL_724772\|202 | ···················· | ·A·················· | ····················· | ···················· | ···················· | ··A················· |
| Y | 7690 hCoV-19/England/LOND-DC52C/2020\|EPI_ISL_724777\|202 | ···················· | ···················· | ····················· | ···················· | ···················· | ···················· |
| Y | 7691 hCoV-19/England/MILK-B87507/2020\|EPI_ISL_724778\|20 | ···················· | ···················· | ····················· | ···················· | ···················· | ···················· |
| Y | 7692 hCoV-19/England/MILK-B875CB/2020\|EPI_ISL_724784\|20 | ···················· | ···················· | ····················· | ···················· | ···················· | ···················· |
| Y | 7693 hCoV-19/England/MILK-B875E9/2020\|EPI_ISL_724785\|20 | ···················· | ···················· | ····················· | ···················· | ···················· | ···················· |
| Y | 7694 hCoV-19/England/MILK-B875F8/2020\|EPI_ISL_724786\|20 | ···················· | ···················· | ····················· | ···················· | ···················· | ···················· |
| Y | 7695 hCoV-19/England/MILK-B87604/2020\|EPI_ISL_724787\|20 | ···················· | ···················· | ····················· | ···················· | ···················· | ···················· |
| Y | 7696 hCoV-19/England/MILK-B87613/2020\|EPI_ISL_724788\|20 | ···················· | ···················· | ····················· | ···················· | ···················· | ···················· |
| Y | 7698 hCoV-19/Japan/FHU-CS24-0227/2020\|EPI_ISL_728206\|20 | ···················· | ···················· | ····················· | ···················· | ···················· | ···················· |
| Y | 7699 hCoV-19/Japan/FHU-CS56-0224/2020\|EPI_ISL_728207\|20 | ···················· | ···················· | ····················· | ···················· | ···················· | ···················· |
| Y | 7700 hCoV-19/Japan/FHU-CS84-0228/2020\|EPI_ISL_728208\|20 | ···················· | ···················· | ····················· | ···················· | ···················· | ···················· |
| Y | 7701 hCoV-19/Malaysia/IMR-WC257440/2020\|EPI_ISL_728209\| | ···················· | ·A·················· | ····················· | ···················· | ···················· | ···················· |
| Y | 7702 hCoV-19/Malaysia/IMR-WC268551/2020\|EPI_ISL_728242\| | ···················· | ·A·················· | ····················· | ···················· | ···················· | ···················· |
| Y | 7703 hCoV-19/Malaysia/IMR-WC268619/2020\|EPI_ISL_728243\| | ···················· | ·A·················· | ····················· | ···················· | ···················· | ···················· |
| Y | 7704 hCoV-19/Malaysia/IMR-WC268890/2020\|EPI_ISL_728244\| | ···················· | ·A·················· | ····················· | ···················· | ···················· | ··A················· |
| Y | 7705 hCoV-19/Malaysia/IMR-WC270611/2020\|EPI_ISL_728245\| | ···················· | ·A·················· | ····················· | ···················· | ···················· | ···················· |
| Y | 7706 hCoV-19/Malaysia/IMR-WC270730/2020\|EPI_ISL_728246\| | ···················· | ·A·················· | ····················· | ···················· | ···················· | ···················· |
| Y | 7707 hCoV-19/Malaysia/IMR-WC272624/2020\|EPI_ISL_728247\| | ···················· | ·A·················· | ····················· | ···················· | ···················· | ···················· |
| Y | 7708 hCoV-19/Malaysia/IMR-WC277466/2020\|EPI_ISL_728249\| | ···················· | ·A·················· | ····················· | ···················· | ···················· | ···················· |
| Y | 7709 hCoV-19/Malaysia/IMR-WC277457/2020\|EPI_ISL_728250\| | ···················· | ·A·················· | ····················· | ···················· | ···················· | ···················· |
| Y | 7710 hCoV-19/Malaysia/IMR-WC280729/2020\|EPI_ISL_728251\| | ···················· | ·A·················· | ····················· | ···················· | ···················· | ···················· |
| Y | 7711 hCoV-19/Malaysia/IMR-WC280757/2020\|EPI_ISL_728252\| | ···················· | ·A·················· | ····················· | ···················· | ···················· | ···················· |
| Y | 7712 hCoV-19/India/MH-BJMC-22757/2020\|EPI_ISL_728254\|20 | ···················· | ···················· | ····················· | ···················· | ···················· | ···················· |
| Y | 7713 hCoV-19/India/MH-BJMC-22259/2020\|EPI_ISL_728255\|20 | ···················· | ···················· | ····················· | ···················· | ···················· | ···················· |
| Y | 7714 hCoV-19/India/MH-BJMC-22244/2020\|EPI_ISL_728256\|20 | ···················· | ···················· | ····················· | ···················· | ···················· | ···················· |
| Y | 7715 hCoV-19/India/MH-BJMC-15009/2020\|EPI_ISL_728257\|20 | ···················· | ···················· | ····················· | ···················· | ···················· | ···················· |
| Y | 7716 hCoV-19/India/MH-BJMC-13585/2020\|EPI_ISL_728258\|20 | ···················· | ···················· | ····················· | ···················· | ···················· | ···················· |
| Y | 7717 hCoV-19/India/MH-BJMC-8677/2020\|EPI_ISL_728259\|202 | ···················· | ···················· | ····················· | ···················· | ···················· | ···················· |
| Y | 7718 hCoV-19/India/MH-BJMC-9070/2020\|EPI_ISL_728260\|202 | ···················· | ···················· | ····················· | ···················· | ···················· | ···················· |
| Y | 7719 hCoV-19/India/MH-BJMC-9069/2020\|EPI_ISL_728261\|202 | ···················· | ···················· | ····················· | ···················· | ···················· | ···················· |
| Y | 7720 hCoV-19/India/MH-BJMC-9065/2020\|EPI_ISL_728262\|202 | ···················· | ···················· | ····················· | ···················· | ···················· | ···················· |
| Y | 7721 hCoV-19/India/MH-BJMC-8860/2020\|EPI_ISL_728263\|202 | ···················· | ···················· | ····················· | ···················· | ···················· | ···················· |
| Y | 7722 hCoV-19/India/MH-BJMC-13308/2020\|EPI_ISL_728264\|20 | ···················· | ···················· | ····················· | ···················· | ···················· | ···················· |
| Y | 7723 hCoV-19/India/MH-BJMC-13327/2020\|EPI_ISL_728265\|20 | ···················· | ···················· | ····················· | ···················· | ···················· | ···················· |
| Y | 7724 hCoV-19/India/MH-BJMC-11016/2020\|EPI_ISL_728266\|20 | ···················· | ···················· | ····················· | ···················· | ···················· | ···················· |
| Y | 7725 hCoV-19/India/MH-BJMC-11034/2020\|EPI_ISL_728267\|20 | ···················· | ···················· | ····················· | ···················· | ···················· | ···················· |
| Y | 7726 hCoV-19/India/MH-BJMC-12267/2020\|EPI_ISL_728268\|20 | ···················· | ···················· | ····················· | ···················· | ···················· | ···················· |
| Y | 7727 hCoV-19/India/MH-BJMC-22738/2020\|EPI_ISL_728269\|20 | ···················· | ···················· | ····················· | ···················· | ···················· | ···················· |
| Y | 7728 hCoV-19/India/MH-BJMC-11052/2020\|EPI_ISL_728270\|20 | ···················· | ···················· | ····················· | ···················· | ···················· | ···················· |
| Y | 7729 hCoV-19/India/MH-BJMC-19749/2020\|EPI_ISL_728271\|20 | ···················· | ···················· | ····················· | ···················· | ···················· | ···················· |
| Y | 7730 hCoV-19/Morocco/FMP-3/2020\|EPI_ISL_728272\|2020-11 | ···················· | ···················· | ····················· | ···················· | ···················· | ···················· |
| Y | 7731 hCoV-19/Morocco/FMP-4/2020\|EPI_ISL_728273\|2020-11 | ···················· | ···················· | ····················· | ···················· | ···················· | ···················· |
| Y | 7732 hCoV-19/Morocco/FMP-5/2020\|EPI_ISL_728274\|2020-11 | ···················· | ···················· | ····················· | ···················· | ···················· | ···················· |
| Y | 7733 hCoV-19/Morocco/FMP-6/2020\|EPI_ISL_728275\|2020-11 | ···················· | ···················· | ····················· | ···················· | ···················· | ···················· |
| Y | 7734 hCoV-19/Morocco/FMP-7/2020\|EPI_ISL_728276\|2020-11 | ···················· | ···················· | ····················· | ···················· | ···················· | ···················· |

| | | | | | | | | |
|---|---|---|---|---|---|---|---|---|
| Y | 7735 hCoV-19/Morocco/FMP-8/2020\|EPI_ISL_728277\|2020-11 | ···················· | ···················· | ···················· | ···················· | ···················· | ···················· | |
| Y | 7736 hCoV-19/Italy/LAZ-INMI-106/2020\|EPI_ISL_728279\|202 | ···················· | ···················· | ···················· | ···················· | ······T············· | ···················· | |
| Y | 7737 hCoV-19/Italy/LAZ-INMI-107/2020\|EPI_ISL_728280\|202 | ···················· | ···················· | ···················· | ···················· | ······T············· | ···················· | |
| Y | 7739 hCoV-19/Italy/LAZ-INMI-109/2020\|EPI_ISL_728282\|202 | ···················· | ····A··············· | ···················· | ···················· | ···················· | ···················· | |
| Y | 7742 hCoV-19/Italy/LAZ-INMI-112/2020\|EPI_ISL_728285\|202 | ···················· | ···················· | ···················· | ···················· | ···················· | ···················· | |
| Y | 7743 hCoV-19/Italy/LAZ-INMI-113/2020\|EPI_ISL_728286\|202 | ···················· | ···················· | ···················· | ···················· | ···················· | ···················· | |
| Y | 7744 hCoV-19/Italy/LAZ-INMI-114/2020\|EPI_ISL_728287\|202 | ···················· | ···················· | ···················· | ···················· | ···················· | ···················· | |
| Y | 7745 hCoV-19/Germany/TH-IIMK-CaSe-3/2020\|EPI_ISL_728288 | ···················· | ···················· | ···················· | ···················· | ···················· | ···················· | |
| Y | 7746 hCoV-19/Morocco/FMP-9/20203\|EPI_ISL_728289\|2020-11 | ···················· | ···················· | ···················· | ···················· | ···················· | ···················· | |
| Y | 7747 hCoV-19/Morocco/FMP-10/20203\|EPI_ISL_728290\|2020-1 | ···················· | ······A············· | ···················· | ···················· | ···················· | ···················· | |
| Y | 7748 hCoV-19/Morocco/FMP-11/2020\|EPI_ISL_728291\|2020-11 | ···················· | ···················· | ···················· | ···················· | ···················· | ···················· | |
| Y | 7749 hCoV-19/Germany/TH-IIMK-CaSe-2/2020\|EPI_ISL_728292 | ···················· | ···················· | ···················· | ···················· | ···················· | ···················· | |
| Y | 7750 hCoV-19/Germany/TH-IIMK-CaSe-4/2020\|EPI_ISL_728293 | ···················· | ···················· | ···················· | ···················· | ···················· | ···················· | |
| Y | 7751 hCoV-19/Germany/TH-IIMK-CaSe-5/2020\|EPI_ISL_728294 | ···················· | ···················· | ···················· | ···················· | ···················· | ···················· | |
| Y | 7752 hCoV-19/Morocco/FMP-12/2020\|EPI_ISL_728295\|2020-11 | ···················· | ···················· | ···················· | ···················· | ···················· | ···················· | |
| Y | 7753 hCoV-19/Germany/TH-IIMK-CaSe-6/2020\|EPI_ISL_728296 | ···················· | ···················· | ···················· | ···················· | ···················· | ···················· | |
| Y | 7754 hCoV-19/Morocco/FMP-13/2020\|EPI_ISL_728297\|2020-11 | ···················· | ···················· | ···················· | ···················· | ···················· | ···················· | |
| Y | 7755 hCoV-19/Germany/TH-IIMK-CaSe-7/2020\|EPI_ISL_728298 | ···················· | ···················· | ···················· | ···················· | ···················· | ···················· | |
| Y | 7756 hCoV-19/England/CAMC-C3F303/2020\|EPI_ISL_728299\|20 | ···················· | ···················· | ···················· | ···················· | ······T············· | ···················· | |
| Y | 7757 hCoV-19/Germany/DE-SH-ChVir3042/2020\|EPI_ISL_72952 | ···················· | ···················· | ···················· | ···················· | ···················· | ···················· | |
| Y | 7758 hCoV-19/Germany/DE-SH-ChVir3045/2020\|EPI_ISL_72952 | ···················· | ···················· | ···················· | ···················· | ···················· | ···················· | |
| Y | 7759 hCoV-19/Germany/DE-SH-ChVir3052/2020\|EPI_ISL_72952 | ···················· | ···················· | ···················· | ···················· | ···················· | ···················· | |
| Y | 7760 hCoV-19/Germany/DE-SH-ChVir3053/2020\|EPI_ISL_72952 | ···················· | ···················· | ···················· | ···················· | ···················· | ···················· | |
| Y | 7761 hCoV-19/Germany/DE-SH-ChVir3054/2020\|EPI_ISL_72952 | ···················· | ···················· | ···················· | ···················· | ···················· | ···················· | |
| Y | 7762 hCoV-19/Germany/DE-SH-ChVir3058/2020\|EPI_ISL_72952 | ···················· | ···················· | ···················· | ···················· | ···················· | ···················· | |
| Y | 7763 hCoV-19/Germany/DE-SH-ChVir3060/2020\|EPI_ISL_72952 | ···················· | ···················· | ···················· | ···················· | ···················· | ···················· | |
| Y | 7764 hCoV-19/Germany/DE-SH-ChVir3061/2020\|EPI_ISL_72952 | ···················· | ···················· | ···················· | ···················· | ···················· | ···················· | |
| Y | 7765 hCoV-19/Germany/DE-SH-ChVir3065/2020\|EPI_ISL_72952 | ···················· | ···················· | ···················· | ···················· | ···················· | ···················· | |
| Y | 7766 hCoV-19/Germany/DE-BE-ChVir3083/2020\|EPI_ISL_72952 | ···················· | ···················· | ···················· | ···················· | ···················· | ···················· | |
| Y | 7767 hCoV-19/Germany/ChVir3098/2020\|EPI_ISL_729530\|2020 | ···················· | ···················· | ···················· | ···················· | ···················· | ···················· | |
| Y | 7768 hCoV-19/Germany/DE-BE-ChVir3100/2020\|EPI_ISL_72953 | ···················· | ···················· | ···················· | ···················· | ···················· | ···················· | |
| Y | 7769 hCoV-19/Germany/DE-BB-ChVir3101/2020\|EPI_ISL_72953 | ···················· | ···················· | ···················· | ···················· | ···················· | ···················· | |
| Y | 7770 hCoV-19/Germany/DE-BE-ChVir3106/2020\|EPI_ISL_72953 | ···················· | ···················· | ···················· | ···················· | ···················· | ···················· | |
| Y | 7771 hCoV-19/Germany/ChVir3137/2020\|EPI_ISL_729534\|2020 | ···················· | ···················· | ···················· | ···················· | ···················· | ···················· | |
| Y | 7772 hCoV-19/Germany/DE-SH-ChVir3184/2020\|EPI_ISL_72953 | ···················· | ···················· | ···················· | ···················· | ···················G | ···················· | 1 |
| Y | 7773 hCoV-19/Germany/DE-SH-ChVir3185/2020\|EPI_ISL_72953 | ···················· | ·············A······ | ···················· | ···················· | ···················· | ···················· | |
| Y | 7774 hCoV-19/Germany/DE-SH-ChVir3187/2020\|EPI_ISL_72953 | ···················· | ···················· | ···················· | ···················· | ···················· | ···················· | |
| Y | 7775 hCoV-19/Germany/DE-SH-ChVir3192/2020\|EPI_ISL_72953 | ···················· | ···················· | ···················· | ···················· | ···················· | ···················· | |
| Y | 7776 hCoV-19/Germany/DE-SH-ChVir3193/2020\|EPI_ISL_72953 | ···················· | ···················· | ···················· | ···················· | ···················· | ···················· | |
| Y | 7777 hCoV-19/Germany/DE-SH-ChVir3218/2020\|EPI_ISL_72954 | ···················· | ···················· | ···················· | ···················· | ···················· | ···················· | |
| Y | 7778 hCoV-19/Germany/DE-SH-ChVir3219/2020\|EPI_ISL_72954 | ···················· | ···················· | ···················· | ···················· | ···················· | ···················· | |
| Y | 7779 hCoV-19/Germany/DE-BB-ChVir3232/2020\|EPI_ISL_72954 | ···················· | ···················· | ···················· | ···················· | ···················· | ···················· | |
| Y | 7780 hCoV-19/Germany/DE-SH-ChVir3031/2020\|EPI_ISL_72954 | ···················· | ···················· | ···················· | ···················· | ···················· | ···················· | |
| Y | 7781 hCoV-19/Germany/ChVir7055/2020\|EPI_ISL_729560\|2020 | ···················· | ···················· | ···················· | ···················· | ···················· | ···················· | |
| Y | 7782 hCoV-19/Germany/DE-BE-ChVir4453/2020\|EPI_ISL_72956 | ···················· | ···················· | ···················· | ···················· | ···················· | ···················· | |
| Y | 7783 hCoV-19/Germany/DE-SH-ChVir8177/2020\|EPI_ISL_72956 | ···················· | ···················· | ···················· | ···················· | ···················· | ···················· | |
| Y | 7784 hCoV-19/Germany/DE-SH-ChVir8179/2020\|EPI_ISL_72956 | ···················· | ···················· | ···················· | ···················· | ···················· | ···················· | |
| Y | 7785 hCoV-19/Germany/DE-SH-ChVir8180/2020\|EPI_ISL_72956 | ···················· | ···················· | ···················· | ···················· | ···················· | ···················· | |
| Y | 7786 hCoV-19/Germany/DE-SH-ChVir8182/2020\|EPI_ISL_72956 | ···················· | ···················· | ···················· | ···················· | ···················· | ···················· | |
| Y | 7787 hCoV-19/Germany/DE-SH-ChVir8183/2020\|EPI_ISL_72956 | ···················· | ···················· | ···················· | ···················· | ···················· | ···················· | |
| Y | 7788 hCoV-19/Germany/DE-SH-ChVir8184/2020\|EPI_ISL_72956 | ···················· | ···················· | ···················· | ···················· | ···················· | ···················· | |
| Y | 7789 hCoV-19/Germany/DE-SH-ChVir8190/2020\|EPI_ISL_72956 | ···················· | ···················· | ···················· | ···················· | ···················· | ···················· | |
| Y | 7790 hCoV-19/Germany/DE-SH-ChVir8195/2020\|EPI_ISL_72956 | ···················· | ···················· | ···················· | ···················· | ···················· | ···················· | |
| Y | 7791 hCoV-19/Germany/DE-SH-ChVir8196/2020\|EPI_ISL_72957 | ···················· | ···················· | ···················· | ···················· | ···················· | ···················· | |
| Y | 7792 hCoV-19/Germany/DE-SH-ChVir8308/2020\|EPI_ISL_72957 | ···················· | ···················· | ···················· | ···················· | ···················· | ···················· | |
| Y | 7793 hCoV-19/Germany/DE-SH-ChVir8310/2020\|EPI_ISL_72957 | ···················· | ···················· | ···················· | ···················· | ···················· | ···················· | |
| Y | 7794 hCoV-19/Germany/DE-SH-ChVir8311/2020\|EPI_ISL_72957 | ···················· | ···················· | ···················· | ···················· | ···················· | ···················· | |
| Y | 7795 hCoV-19/Germany/DE-SH-ChVir8315/2020\|EPI_ISL_72957 | ···················· | ···················· | ···················· | ···················· | ···················· | ···················· | |
| Y | 7796 hCoV-19/Germany/DE-SH-ChVir8317/2020\|EPI_ISL_72957 | ···················· | ···················· | ···················· | ···················· | ···················· | ···················· | |
| Y | 7797 hCoV-19/Germany/DE-SH-ChVir8318/2020\|EPI_ISL_72957 | ···················· | ···················· | ···················· | ···················· | ···················· | ···················· | |
| Y | 7798 hCoV-19/Germany/DE-SH-ChVir8319/2020\|EPI_ISL_72957 | ···················· | ···················· | ···················· | ···················· | ···················· | ···················· | |
| Y | 7799 hCoV-19/Germany/DE-SH-ChVir8321/2020\|EPI_ISL_72957 | ···················· | ·A·················· | ···········T········ | ···················· | ···················· | ···················· | |
| Y | 7800 hCoV-19/Germany/DE-SH-ChVir8322/2020\|EPI_ISL_72957 | ···················· | ·A·················· | ···········T········ | ···················· | ···················· | ···················· | |
| Y | 7801 hCoV-19/Germany/DE-SH-ChVir8325/2020\|EPI_ISL_72958 | ···················· | ···················· | ···················· | ···················· | ···················· | ···················· | |
| Y | 7802 hCoV-19/Germany/DE-SH-ChVir8326/2020\|EPI_ISL_72958 | ···················· | ···················· | ···················· | ···················· | ···················· | ···················· | |
| Y | 7803 hCoV-19/Germany/DE-SH-ChVir8327/2020\|EPI_ISL_72958 | ···················· | ·A·················· | ···········T········ | ···················· | ···················· | ···················· | |
| Y | 7804 hCoV-19/Germany/DE-SH-ChVir8328/2020\|EPI_ISL_72958 | ···················· | ···················· | ···················· | ···················· | ···················· | ···················· | |
| Y | 7805 hCoV-19/Germany/DE-BE-ChVir8329/2020\|EPI_ISL_72958 | ···················· | ···················· | ···················· | ···················· | ···················· | ···················· | |
| Y | 7806 hCoV-19/Germany/DE-SH-ChVir8332/2020\|EPI_ISL_72958 | ···················· | ···················· | ···················· | ···················· | ···················· | ···················· | |
| Y | 7807 hCoV-19/Germany/DE-SH-ChVir8333/2020\|EPI_ISL_72958 | ···················· | ···················· | ···················· | ···················· | ···················· | ···················· | |
| Y | 7808 hCoV-19/Germany/DE-SH-ChVir8334/2020\|EPI_ISL_72958 | ···················· | ···················· | ···················· | ···················· | ···················· | ···················· | |
| Y | 7809 hCoV-19/Germany/DE-SH-ChVir8587/2020\|EPI_ISL_72958 | ···················· | ···················· | ···················· | ···················· | ····T··············· | ···················· | |
| Y | 7810 hCoV-19/Germany/DE-SH-ChVir8595/2020\|EPI_ISL_72958 | ···················· | ···················· | ···················· | ···················· | ···················· | ···················· | |
| Y | 7811 hCoV-19/Germany/DE-SH-ChVir8596/2020\|EPI_ISL_72959 | ···················· | ···················· | ···················· | ···················· | ···················· | ···················· | |
| Y | 7812 hCoV-19/Germany/DE-SH-ChVir8599/2020\|EPI_ISL_72959 | ···················· | ···················· | ···················· | ···················· | ···················· | ···················· | |
| Y | 7813 hCoV-19/Germany/DE-SH-ChVir8604/2020\|EPI_ISL_72959 | ···················· | ···················· | ···················· | ···················· | ···················· | ···················· | |
| Y | 7814 hCoV-19/Germany/DE-SH-ChVir8606/2020\|EPI_ISL_72959 | ···················· | ···················· | ···················· | ···················· | ···················· | ···················· | |
| Y | 7815 hCoV-19/Germany/DE-SH-ChVir8607/2020\|EPI_ISL_72959 | ···················· | ···················· | ···················· | ···················· | ···················· | ···················· | |
| Y | 7816 hCoV-19/Germany/DE-SH-ChVir8608/2020\|EPI_ISL_72959 | ···················· | ···················· | ···················· | ···················· | ···················· | ···················· | |
| Y | 7817 hCoV-19/Germany/ChVir8634/2020\|EPI_ISL_729596\|2020 | ···················· | ···················· | ···················· | ···················· | ···················· | ···················· | |

```
Y 7818 hCoV-19/Germany/ChVir8637/2020|EPI_ISL_729597|2020 ···················· ···················· ···················· ···················· ···················· ····················
Y 7819 hCoV-19/Germany/ChVir8967/2020|EPI_ISL_729598|2020 ···················· ···················· ···················· ···················· ······T············· ····················
Y 7820 hCoV-19/Germany/DE-BE-ChVir8988/2020|EPI_ISL_72959 ···················· ···················· ···················· ···················· ···················· ····················
Y 7821 hCoV-19/Jordan/AQ-ALSR-5263/2020|EPI_ISL_730501|20 ···················· ···················· ···················· ···················· ···················· ····················
Y 7822 hCoV-19/Jordan/AQ-ALSR-5264/2020|EPI_ISL_730502|20 ···················· ···················· ···················· ···················· ···················· ····················
Y 7823 hCoV-19/Jordan/AQ-ALSR-5265/2020|EPI_ISL_730503|20 ···················· ···················· ···················· ···················· ···················· ····················
Y 7824 hCoV-19/Jordan/AQ-ALSR-5266/2020|EPI_ISL_730504|20 ···················· ···················· ···················· ···················· ···················· ····················
Y 7825 hCoV-19/Jordan/AQ-ALSR-5267/2020|EPI_ISL_730505|20 ···················· ···················· ···················· ···················· ···················· ····················
Y 7826 hCoV-19/Jordan/AQ-ALSR-5268/2020|EPI_ISL_730506|20 ···················· ·A·················· ···················· ···················· ···················· ··A·················
Y 7827 hCoV-19/Jordan/AQ-ALSR-5269/2020|EPI_ISL_730507|20 ···················· ·A·················· ···················· ···················· ···················· ··A·················
Y 7828 hCoV-19/Jordan/AQ-ALSR-5270/2020|EPI_ISL_730508|20 ···················· ···················· ···················· ···················· ···················· ····················
Y 7829 hCoV-19/Jordan/AQ-ALSR-5271/2020|EPI_ISL_730509|20 ···················· ···················· ···················· ···················· ···················· ····················
Y 7830 hCoV-19/Jordan/AQ-ALSR-5272/2020|EPI_ISL_730510|20 ···················· ···················· ···················· ···················· ···················· ····················
Y 7831 hCoV-19/Jordan/AQ-ALSR-5273/2020|EPI_ISL_730511|20 ···················· ···················· ···················· ···················· ···················· ····················
Y 7832 hCoV-19/Jordan/AQ-ALSR-5274/2020|EPI_ISL_730512|20 ···················· ···················· ···················· ···················· ···················· ····················
Y 7833 hCoV-19/Jordan/AQ-ALSR-5275/2020|EPI_ISL_730513|20 ···················· ···················· ···················· ···················· ···················· ····················
Y 7834 hCoV-19/Jordan/AQ-ALSR-5276/2020|EPI_ISL_730514|20 ···················· ···················· ···················· ···················· ···················· ····················
Y 7835 hCoV-19/Jordan/AQ-ALSR-5277/2020|EPI_ISL_730515|20 ···················· ···················· ···················· ···················· ···················· ····················
Y 7836 hCoV-19/Jordan/AQ-ALSR-5278/2020|EPI_ISL_730516|20 ···················· ···················· ···················· ···················· ···················· ····················
Y 7837 hCoV-19/Jordan/IR-ALSR-5279/2020|EPI_ISL_730517|20 ···················· ···················· ···················· ···················· ········T··········· ····················
Y 7838 hCoV-19/Jordan/AM-ALSR-5280/2020|EPI_ISL_730518|20 ···················· ···················· ···················· ···················· ········T··········· ····················
Y 7839 hCoV-19/Jordan/AM-ALSR-5281/2020|EPI_ISL_730519|20 ···················· ···················· ···················· ···················· ···················· ····················
Y 7840 hCoV-19/Jordan/AM-ALSR-5283/2020|EPI_ISL_730520|20 ···················· ·A·················· ···················· ···················· ···················· ··A·················
Y 7841 hCoV-19/Jordan/AM-ALSR-5284/2020|EPI_ISL_730521|20 ···················· ·A·················· ···················· ···················· ···················· ··A·················
Y 7842 hCoV-19/Jordan/AM-ALSR-5285/2020|EPI_ISL_730522|20 ···················· ···················· ···················· ···················· ···················· ····················
Y 7843 hCoV-19/Jordan/AM-ALSR-5294/2020|EPI_ISL_730531|20 ···················· ···················· ···················· ···················· ···················· ····················
Y 7844 hCoV-19/Jordan/AM-ALSR-5310/2020|EPI_ISL_730546|20 ···················· ···················· ···················· ···················· ···················· ····················
Y 7845 hCoV-19/Jordan/AM-ALSR-5312/2020|EPI_ISL_730548|20 ···················· ···················· ···················· ···················· ···················· ····················
Y 7846 hCoV-19/Jordan/AM-ALSR-5313/2020|EPI_ISL_730549|20 ···················· ·A·················· ···················· ···················· ···················· ··A·················
Y 7847 hCoV-19/Jordan/AM-ALSR-5314/2020|EPI_ISL_730550|20 ···················· ·A·················· ···················· ···················· ···················· ··A·················
Y 7848 hCoV-19/Jordan/AM-ALSR-5315/2020|EPI_ISL_730551|20 ···················· ···················· ···················· ···················· ···················· ····················
Y 7849 hCoV-19/Jordan/AM-ALSR-5316/2020|EPI_ISL_730552|20 ···················· ···················· ···················· ···················· ···················· ····················
Y 7850 hCoV-19/Jordan/AM-ALSR-5317/2020|EPI_ISL_730553|20 ···················· ···················· ···················· ···················· ···················· ····················
Y 7851 hCoV-19/Jordan/AM-ALSR-5318/2020|EPI_ISL_730554|20 ···················· ···················· ···················· ···················· ···················· ····················
Y 7852 hCoV-19/Jordan/AM-ALSR-5319/2020|EPI_ISL_730555|20 ···················· ···················· ···················· ···················· ···················· ····················
Y 7853 hCoV-19/Jordan/AM-ALSR-5320/2020|EPI_ISL_730556|20 ···················· ·A·················· ···················· ···················· ···················· ··A·················
Y 7854 hCoV-19/Jordan/AM-ALSR-5321/2020|EPI_ISL_730557|20 ···················· ···················· ···················· ···················· ···················· ····················
Y 7855 hCoV-19/Jordan/AM-ALSR-5322/2020|EPI_ISL_730558|20 ···················· ···················· ···················· ···················· ···················· ····················
Y 7856 hCoV-19/Jordan/AM-ALSR-5323/2020|EPI_ISL_730559|20 ···················· ·A·················· ···················· ···················· ···················· ··A·················
Y 7857 hCoV-19/Jordan/AM-ALSR-5324/2020|EPI_ISL_730560|20 ···················· ·A·················· ···················· ···················· ···················· ··A·················
Y 7858 hCoV-19/Jordan/AM-ALSR-5325/2020|EPI_ISL_730561|20 ···················· ···················· ···················· ···················· ···················· ····················
Y 7859 hCoV-19/Jordan/AM-ALSR-5326/2020|EPI_ISL_730562|20 ···················· ···················· ···················· ···················· ······T············· ····················
Y 7860 hCoV-19/Jordan/AM-ALSR-5327/2020|EPI_ISL_730563|20 ···················· ···················· ···················· ···················· ···················· ····················
Y 7861 hCoV-19/USA/MI-UM-10036869928/2020|EPI_ISL_730565| ···················· ···················· ···················· ···················· ···················· ····················
Y 7863 hCoV-19/Turkey/GUMV_36208/2020|EPI_ISL_730567|2020 ···················· ···················· ···················· ···················· ···················· ····················
Y 7864 hCoV-19/Turkey/Ankara_GUMV_25210/2020|EPI_ISL_7305 ···················· ···················· ···················· ···················· ···················· ····················
Y 7865 hCoV-19/Turkey/Ankara_GUMV_37230/2020|EPI_ISL_7305 ···················· ···················· ···················· ···················· ···················· ····················
Y 7866 hCoV-19/Turkey/Ankara_GUMV_22731/2020|EPI_ISL_7305 ···················· ···················· ···················· ···················· ···················· ····················
Y 7867 hCoV-19/Turkey/Ankara_GUMV_28458/2020|EPI_ISL_7305 ···················· ···················· ···················· ···················· ···················· ····················
Y 7868 hCoV-19/Turkey/Ankara_GUMV_18121/2020|EPI_ISL_7305 ···················· ···················· ···················· ···················· ···················· ····················
Y 7869 hCoV-19/Turkey/Ankara_33508/2020|EPI_ISL_730575|20 ···················· ···················· ···················· ···················· ···················· ····················
Y 7870 hCoV-19/Turkey/Ankara_GUMV_10711/2020|EPI_ISL_7305 ···················· ···················· ···················· ···················· ···················· ····················
Y 7871 hCoV-19/Turkey/Ankara_GUMV_42541/2020|EPI_ISL_7305 ···················· ·A·················· ···················· ···················· ···················· ··A·················
Y 7872 hCoV-19/England/ALDP-C3CD17/2020|EPI_ISL_731832|20 ···················· ···················· ···················· ···················· ···················· ····················
Y 7873 hCoV-19/England/ALDP-C3D790/2020|EPI_ISL_731833|20 ···················· ···················· ···················· ···················· ···················· ····················
Y 7874 hCoV-19/England/ALDP-C3D693/2020|EPI_ISL_731834|20 ···················· ···················· ···················· ···················· ···················· ····················
Y 7875 hCoV-19/England/ALDP-C3C9B9/2020|EPI_ISL_731836|20 ···················· ···················· ···················· ···················· ···················· ····················
Y 7876 hCoV-19/England/ALDP-C3BE60/2020|EPI_ISL_731837|20 ···················· ···················· ···················· ···················· ···················· ····················
Y 7877 hCoV-19/England/ALDP-C3B5D4/2020|EPI_ISL_731838|20 ···················· ···················· ···················· ···················· ···················· ····················
Y 7878 hCoV-19/England/ALDP-C3CB59/2020|EPI_ISL_731839|20 ···················· ···················· ···················· ···················· ···················· ····················
Y 7879 hCoV-19/England/ALDP-C3B255/2020|EPI_ISL_731840|20 ···················· ·A·················· ···················· ···················· ···················· ··A·················
Y 7880 hCoV-19/England/ALDP-C3AB00/2020|EPI_ISL_731841|20 ···················· ···················· ···················· ···················· ···················· ····················
Y 7881 hCoV-19/England/ALDP-C3D736/2020|EPI_ISL_731843|20 ···················· ···················· ···················· ···················· ···················· ····················
Y 7882 hCoV-19/England/ALDP-C3D1B0/2020|EPI_ISL_731845|20 ···················· ···················· ···················· ···················· ···················· ····················
Y 7883 hCoV-19/England/ALDP-C3B02E/2020|EPI_ISL_731846|20 ·········T·········· ···················· ···················· ···················· ···················· ····················
Y 7884 hCoV-19/England/ALDP-C3CBFF/2020|EPI_ISL_731848|20 ···················· ·········T·········· ···················· ···················· ···················· ····················
Y 7885 hCoV-19/England/ALDP-C3DD07/2020|EPI_ISL_731849|20 ···················· ···················· ···················· ···················· ···················· ····················
Y 7886 hCoV-19/England/ALDP-C3BFA9/2020|EPI_ISL_731853|20 ···················· ·A·················· ···················· ···················· ···················· ··A·················
Y 7887 hCoV-19/England/ALDP-C3DCEC/2020|EPI_ISL_731862|20 ···················· ···················· ···················· ···················· ···················· ····················
Y 7888 hCoV-19/England/ALDP-C3D01D/2020|EPI_ISL_731876|20 ···················· ···················· ···················· ···················· ···················· ····················
Y 7889 hCoV-19/England/ALDP-C3CC1A/2020|EPI_ISL_731879|20 ···················· ···················· ···················· ···················· ···················· ····················
Y 7890 hCoV-19/England/ALDP-C3C861/2020|EPI_ISL_731880|20 ···················· ·A·················· ···················· ···················· ···················· ··A·················
Y 7891 hCoV-19/England/ALDP-C3B4F5/2020|EPI_ISL_731881|20 ···················· ···················· ···················· ···················· ···················· ····················
Y 7892 hCoV-19/England/ALDP-C3DCDD/2020|EPI_ISL_731882|20 ···················· ···················· ···················· ···················· ···················· ····················
Y 7893 hCoV-19/England/ALDP-C3BDEB/2020|EPI_ISL_731883|20 ···················· ···················· ···················· ···················· ···················· ····················
Y 7894 hCoV-19/England/ALDP-C3B4E6/2020|EPI_ISL_731884|20 ···················· ···················· ···················· ···················· ···················· ····················
Y 7895 hCoV-19/England/ALDP-C3CDCC/2020|EPI_ISL_731886|20 ···················· ···················· ···················· ···················· ···················· ····················
Y 7896 hCoV-19/England/ALDP-C3B291/2020|EPI_ISL_731887|20 ···················· ···················· ···················· ···················· ···················· ····················
Y 7897 hCoV-19/England/ALDP-C3B0E2/2020|EPI_ISL_731888|20 ···················· ···················· ···················· ···················· ···················· ····················
Y 7898 hCoV-19/England/ALDP-C3D38D/2020|EPI_ISL_731890|20 ···················· ···················· ···················· ···················· ···················· ····················
```

| | | | | | | | | |
|---|---|---|---|---|---|---|---|---|
| Y | 7899 hCoV-19/England/MILK-BFFA74/2020\|EPI_ISL_731892\|20 | .................... | .A.................. | .................... | .................... | .................... | ..A................. | |
| Y | 7900 hCoV-19/England/QEUH-BA3945/2020\|EPI_ISL_731894\|20 | .................... | .................... | .................... | .................... | .................... | .................... | |
| Y | 7901 hCoV-19/England/QEUH-C34605/2020\|EPI_ISL_734101\|20 | .................... | .................... | .................... | .................... | .................... | .................... | |
| Y | 7902 hCoV-19/England/QEUH-C33EA4/2020\|EPI_ISL_734102\|20 | .................... | .................... | .................... | .................... | .................... | .................... | |
| Y | 7903 hCoV-19/England/QEUH-C343E0/2020\|EPI_ISL_734103\|20 | .................... | .................... | .................... | .................... | .................... | .................... | |
| Y | 7904 hCoV-19/England/QEUH-C32E96/2020\|EPI_ISL_734104\|20 | .................... | .................... | .................... | .................... | .................... | .................... | |
| Y | 7905 hCoV-19/England/QEUH-C342E3/2020\|EPI_ISL_734105\|20 | .................... | .A.................. | .................... | .................... | .................... | ..A................. | |
| Y | 7906 hCoV-19/England/QEUH-C34632/2020\|EPI_ISL_734106\|20 | .................... | .................... | .................... | .................... | .................... | .................... | |
| Y | 7907 hCoV-19/England/QEUH-C331D8/2020\|EPI_ISL_734108\|20 | .................... | .................... | .................... | .................... | .................... | .................... | |
| Y | 7908 hCoV-19/Brazil/SP-792/2020\|EPI_ISL_735429\|2020-07- | .................... | .................... | .................... | .................... | .................... | .................... | |
| Y | 7909 hCoV-19/Brazil/SP-793/2020\|EPI_ISL_735430\|2020-07- | .................... | .................... | .................... | .................... | .................... | .................... | |
| Y | 7910 hCoV-19/Brazil/SP-794/2020\|EPI_ISL_735431\|2020-07- | .................... | .................... | .................... | .................... | .................... | .................... | |
| Y | 7911 hCoV-19/Brazil/SP-795/2020\|EPI_ISL_735432\|2020-07- | .................... | .................... | .................... | .................... | .................... | .................... | |
| Y | 7912 hCoV-19/Brazil/SP-796/2020\|EPI_ISL_735433\|2020-07- | .................... | .................... | .................... | .................... | .................... | .................... | |
| Y | 7913 hCoV-19/Rwanda/NRLNAT1001/2020\|EPI_ISL_735436\|2020 | .................... | .................... | .................... | .................... | .................... | .................... | |
| Y | 7914 hCoV-19/Rwanda/NRLNAT1003/2020\|EPI_ISL_735437\|2020 | .................... | .................... | .................... | .................... | .................... | .................... | |
| Y | 7915 hCoV-19/Rwanda/1002/2020\|EPI_ISL_735438\|2020-08-30 | .................... | .................... | .................... | .................... | .................... | .................... | |
| Y | 7916 hCoV-19/England/QEUH-C4BE7C/2020\|EPI_ISL_736700\|20 | .................... | .................... | .................... | .................... | .................... | .................... | |
| Y | 7917 hCoV-19/England/ALDP-C48510/2020\|EPI_ISL_736701\|20 | .................... | .................... | .................... | .................... | .................... | .................... | |
| Y | 7918 hCoV-19/England/MILK-C504EB/2020\|EPI_ISL_736703\|20 | .................... | .................... | .................... | .................... | .................... | .................... | |
| Y | 7919 hCoV-19/England/ALDP-C48C1B/2020\|EPI_ISL_736704\|20 | .................... | .A.................. | .................... | .................... | .................... | .................... | |
| Y | 7920 hCoV-19/England/ALDP-C4852F/2020\|EPI_ISL_736705\|20 | .................... | .................... | .................... | .................... | .................... | .................... | |
| Y | 7921 hCoV-19/England/ALDP-C48BA5/2020\|EPI_ISL_736707\|20 | .................... | .................... | .................... | .................... | .................... | .................... | |
| Y | 7922 hCoV-19/England/ALDP-C48950/2020\|EPI_ISL_736708\|20 | .................... | .................... | .................... | .................... | .................... | .................... | |
| Y | 7923 hCoV-19/England/ALDP-C489C9/2020\|EPI_ISL_736710\|20 | .................... | .................... | .................... | .................... | .................... | .................... | |
| Y | 7924 hCoV-19/England/MILK-C5075B/2020\|EPI_ISL_736713\|20 | .................... | .................... | .................... | .................... | .................... | .................... | |
| Y | 7925 hCoV-19/England/MILK-C506C7/2020\|EPI_ISL_736716\|20 | .................... | .................... | .................... | .................... | .................... | .................... | |
| Y | 7926 hCoV-19/England/ALDP-C48783/2020\|EPI_ISL_736717\|20 | .................... | .................... | .................... | .................... | ......T............. | .................... | |
| Y | 7927 hCoV-19/England/ALDP-C487ED/2020\|EPI_ISL_736718\|20 | .................... | .................... | .................... | .................... | .................... | .................... | |
| Y | 7928 hCoV-19/England/ALDP-C484C8/2020\|EPI_ISL_736720\|20 | .................... | .................... | .................... | .................... | .................... | .................... | |
| Y | 7929 hCoV-19/England/QEUH-C4C224/2020\|EPI_ISL_736721\|20 | .................... | .................... | .................... | .................... | .................... | .................... | |
| Y | 7930 hCoV-19/England/ALDP-C48E15/2020\|EPI_ISL_736726\|20 | .................... | .................... | .................... | .................... | .................... | .................... | |
| Y | 7931 hCoV-19/England/ALDP-C3D226/2020\|EPI_ISL_736760\|20 | .................... | .................... | .................... | .................... | .................... | .................... | |
| Y | 7932 hCoV-19/England/ALDP-C3D1DE/2020\|EPI_ISL_736762\|20 | .................... | .................... | .................... | .................... | .................... | .................... | |
| Y | 7933 hCoV-19/England/ALDP-C3D183/2020\|EPI_ISL_736765\|20 | .................... | .................... | .................... | .................... | .................... | .................... | |
| Y | 7934 hCoV-19/England/ALDP-C3D305/2020\|EPI_ISL_736766\|20 | .................... | .................... | .................... | .................... | .................... | .................... | |
| Y | 7935 hCoV-19/England/ALDP-C3BCC0/2020\|EPI_ISL_736768\|20 | .................... | .................... | .................... | .................... | .................... | .................... | |
| Y | 7936 hCoV-19/England/ALDP-C3D2CC/2020\|EPI_ISL_736769\|20 | .................... | .................... | .................... | .................... | .................... | .................... | |
| Y | 7937 hCoV-19/England/LOND-DB280/2020\|EPI_ISL_724750\|202 | ..................T. | .................... | .................... | .................... | .................... | .................... | 1 |
| Y | 7938 hCoV-19/England/LOND-DB29F/2020\|EPI_ISL_724751\|202 | .................... | .................... | .................... | .................... | .................... | .................... | |
| Y | 7939 hCoV-19/England/LOND-DB2CC/2020\|EPI_ISL_724752\|202 | .................... | .................... | .................... | .................... | .................... | .................... | |
| Y | 7940 hCoV-19/England/LOND-DB350/2020\|EPI_ISL_724759\|202 | .................... | .................... | .................... | .................... | .................... | .................... | |
| Y | 7941 hCoV-19/England/LOND-DB37E/2020\|EPI_ISL_724760\|202 | .................... | .................... | .................... | .................... | .................... | .................... | |
| Y | 7942 hCoV-19/England/LOND-DB46C/2020\|EPI_ISL_724769\|202 | .................... | .................... | .................... | .................... | .................... | .................... | |
| Y | 7943 hCoV-19/England/MILK-B87534/2020\|EPI_ISL_724780\|20 | .................... | .................... | .................... | .................... | .................... | .................... | |
| Y | 7944 hCoV-19/England/MILK-B87543/2020\|EPI_ISL_724781\|20 | .................... | .................... | .................... | .................... | .................... | .................... | |
| Y | 7945 hCoV-19/England/MILK-B87552/2020\|EPI_ISL_724782\|20 | .................... | .................... | .................... | .................... | .................... | .................... | |
| Y | 7946 hCoV-19/England/MILK-B8759E/2020\|EPI_ISL_724783\|20 | .................... | .................... | .................... | .................... | .................... | .................... | |
| Y | 7947 hCoV-19/England/MILK-B8766E/2020\|EPI_ISL_724790\|20 | .................... | .................... | .................... | .................... | .................... | .................... | |
| Y | 7948 hCoV-19/England/MILK-B8768C/2020\|EPI_ISL_724791\|20 | .................... | .................... | .................... | .................... | .................... | .................... | |
| Y | 7949 hCoV-19/England/MILK-B876E6/2020\|EPI_ISL_724792\|20 | .................... | .................... | .................... | .................... | .................... | .................... | |
| Y | 7950 hCoV-19/England/MILK-B876F5/2020\|EPI_ISL_724793\|20 | .................... | .................... | .................... | .................... | .................... | .................... | |
| Y | 7951 hCoV-19/England/MILK-B87710/2020\|EPI_ISL_724794\|20 | .................... | .................... | .................... | .................... | .................... | .................... | |
| Y | 7952 hCoV-19/England/MILK-B8772F/2020\|EPI_ISL_724795\|20 | .................... | .................... | .................... | .................... | .................... | .................... | |
| Y | 7953 hCoV-19/England/MILK-B8774D/2020\|EPI_ISL_724796\|20 | .................... | .................... | .................... | .................... | .................... | .................... | |
| Y | 7954 hCoV-19/England/MILK-B877C5/2020\|EPI_ISL_724799\|20 | .................... | .................... | .................... | .................... | .................... | .................... | |
| Y | 7955 hCoV-19/England/QEUH-B9F065/2020\|EPI_ISL_728300\|20 | .................... | .................... | .................... | .................... | .................... | .................... | |
| Y | 7956 hCoV-19/Morocco/FMP-14/2020\|EPI_ISL_728301\|2020-11 | .................... | .................... | .................... | .................... | .................... | .................... | |
| Y | 7957 hCoV-19/Germany/TH-IIMK-CaSe-8/2020\|EPI_ISL_728302 | .................... | .................... | .................... | .................... | .................... | .................... | |
| Y | 7958 hCoV-19/Germany/TH-IIMK-CaSe-9/2020\|EPI_ISL_728303 | .................... | .................... | .................... | .................... | .................... | .................... | |
| Y | 7960 hCoV-19/France/ARA-105681/2020\|EPI_ISL_728305\|2020 | .................... | .................... | .................... | .................... | .................... | .................... | |
| Y | 7961 hCoV-19/France/ARA-105758/2020\|EPI_ISL_728306\|2020 | .................... | .................... | .................... | .................... | .................... | .................... | |
| Y | 7962 hCoV-19/France/ARA-105851/2020\|EPI_ISL_728307\|2020 | .................... | .................... | .................... | .................... | NNN................. | .................... | 1 |
| Y | 7963 hCoV-19/France/ARA-106294/2020\|EPI_ISL_728308\|2020 | .................... | .................... | .................... | .................... | .................... | .................... | |
| Y | 7964 hCoV-19/France/ARA-106297/2020\|EPI_ISL_728309\|2020 | .................... | .................... | .................... | .................... | .................... | .................... | |
| Y | 7965 hCoV-19/Germany/DE-SH-ChVir9131/2020\|EPI_ISL_72961 | .................... | ...A................ | .................... | .................... | .................... | .................... | |
| Y | 7966 hCoV-19/Germany/DE-SH-ChVir9277/2020\|EPI_ISL_72961 | .................... | .A.................. | .................... | .................... | .................... | .................... | |
| Y | 7967 hCoV-19/Germany/DE-SH-ChVir9280/2020\|EPI_ISL_72961 | .................... | .................... | .................... | .................... | ......T............. | .................... | |
| Y | 7968 hCoV-19/Germany/DE-SH-ChVir9286/2020\|EPI_ISL_72961 | .................... | .................... | .................... | .................... | .................... | .................... | |
| Y | 7969 hCoV-19/Germany/DE-SH-ChVir9288/2020\|EPI_ISL_72961 | .................... | .................... | .................... | .................... | .................... | .................... | |
| Y | 7970 hCoV-19/Germany/DE-SH-ChVir9274/2020\|EPI_ISL_72961 | .................... | .................... | .................... | .................... | .................... | .................... | |
| Y | 7971 hCoV-19/Germany/DE-SH-ChVir9275/2020\|EPI_ISL_72961 | .................... | .................... | .................... | .................... | .................... | .................... | |
| Y | 7972 hCoV-19/Germany/DE-SH-ChVir9284/2020\|EPI_ISL_72961 | .................... | .................... | .................... | .................... | .................... | .................... | |
| Y | 7973 hCoV-19/Germany/DE-SH-ChVir9287/2020\|EPI_ISL_72961 | .................... | .................... | .................... | .................... | .................... | .................... | |
| Y | 7974 hCoV-19/Germany/DE-SH-ChVir9636/2020\|EPI_ISL_72961 | .................... | .................... | .................... | .................... | .................... | .................... | |
| Y | 7975 hCoV-19/Germany/DE-SH-ChVir9932/2020\|EPI_ISL_72962 | .................... | .................... | .................... | .................... | .................... | .................... | |
| Y | 7976 hCoV-19/Germany/DE-SH-ChVir9935/2020\|EPI_ISL_72962 | .................... | .................... | .................... | .................... | .................... | .................... | |
| Y | 7977 hCoV-19/Germany/DE-SH-ChVir9946/2020\|EPI_ISL_72962 | .................... | .A.................. | .................... | .................... | .................... | .................... | |
| Y | 7978 hCoV-19/Germany/DE-SH-ChVir9947/2020\|EPI_ISL_72962 | .................... | .................... | .................... | .................... | .................... | .................... | |
| Y | 7979 hCoV-19/Germany/DE-SH-ChVir9995/2020\|EPI_ISL_72962 | .................... | .................... | .................... | .................... | .................... | .................... | |

| | | | | | | | | |
|---|---|---|---|---|---|---|---|---|
| Y | 7980 | hCoV-19/Germany/DE-SH-ChVir20001/2020\|EPI_ISL_7296 | ···················· | ···················· | ····················· | ···················· | ···················· | ···················· |
| Y | 7981 | hCoV-19/Germany/DE-SH-ChVir20002/2020\|EPI_ISL_7296 | ···················· | ···················· | ····················· | ···················· | ···················· | ···················· |
| Y | 7982 | hCoV-19/Germany/DE-SH-ChVir20005/2020\|EPI_ISL_7296 | ···················· | ···················· | ····················· | ···················· | ···················· | ···················· |
| Y | 7983 | hCoV-19/Germany/DE-NW-ChVir20006/2020\|EPI_ISL_7296 | ···················· | ···A················ | ····················· | ···················· | ···················· | ···················· |
| Y | 7984 | hCoV-19/Germany/DE-SH-ChVir20012/2020\|EPI_ISL_7296 | ···················· | ···················· | ····················· | ···················· | ···················· | ···················· |
| Y | 7985 | hCoV-19/Germany/DE-SH-ChVir20031/2020\|EPI_ISL_7296 | ···················· | ···················· | ····················· | ···················· | ···················· | ···················· |
| Y | 7986 | hCoV-19/Germany/DE-SH-ChVir20036/2020\|EPI_ISL_7296 | ···················· | ···A················ | ····················· | ···················· | ···················· | ···················· |
| Y | 7987 | hCoV-19/Germany/DE-SH-ChVir20042/2020\|EPI_ISL_7296 | ···················· | ···················· | ····················· | ···················· | ···················· | ···················· |
| Y | 7988 | hCoV-19/Germany/DE-SH-ChVir20043/2020\|EPI_ISL_7296 | ···················· | ···················· | ····················· | ···················· | ···················· | ···················· |
| Y | 7989 | hCoV-19/Germany/DE-SH-ChVir20049/2020\|EPI_ISL_7296 | ···················· | ···················· | ····················· | ···················· | ···················· | ···················· |
| Y | 7990 | hCoV-19/Germany/DE-SH-ChVir20296/2020\|EPI_ISL_7296 | ···················· | ···················· | ····················· | ···················· | ···················· | ···················· |
| Y | 7991 | hCoV-19/Germany/DE-SH-ChVir20297/2020\|EPI_ISL_7296 | ···················· | ···················· | ····················· | ···················· | ···················· | ···················· |
| Y | 7992 | hCoV-19/Germany/DE-SH-ChVir20299/2020\|EPI_ISL_7296 | ···················· | ···················· | ····················· | ···················· | ···················· | ···················· |
| Y | 7993 | hCoV-19/Germany/DE-SH-ChVir20300/2020\|EPI_ISL_7296 | ···················· | ···················· | ····················· | ···················· | ···················· | ···················· |
| Y | 7994 | hCoV-19/Germany/DE-SH-ChVir20301/2020\|EPI_ISL_7296 | ···················· | ···················· | ····················· | ···················· | ···················· | ···················· |
| Y | 7995 | hCoV-19/Turkey/Ankara_GUMV_41954/2020\|EPI_ISL_7306 | ···················· | ···················· | ····················· | ···················· | ···················· | ···················· |
| Y | 7996 | hCoV-19/Switzerland/AG-UZH-IMV122/2020\|EPI_ISL_730 | ···················· | ···················· | ····················· | ···················· | ···················· | ···················· |
| Y | 7997 | hCoV-19/Portugal/PT1777/2020\|EPI_ISL_731908\|2020-1 | ···················· | ···················· | ····················· | ···················· | ···················· | ···················· |
| Y | 7998 | hCoV-19/Portugal/PT1757/2020\|EPI_ISL_731909\|2020-1 | ···················· | ···················· | ····················· | ···················· | ···················· | ···················· |
| Y | 7999 | hCoV-19/Portugal/PT1680/2020\|EPI_ISL_731917\|2020-1 | ···················· | ···················· | ····················· | ···················· | ···················· | ···················· |
| Y | 8000 | hCoV-19/Portugal/PT1642/2020\|EPI_ISL_731918\|2020-1 | ···················· | ···················· | ····················· | ···················· | ···················· | ···················· |
| Y | 8001 | hCoV-19/Portugal/PT1643/2020\|EPI_ISL_731919\|2020-1 | ···················· | ···················· | ····················· | ···················· | ···················· | ···················· |
| Y | 8002 | hCoV-19/Portugal/PT1645/2020\|EPI_ISL_731920\|2020-1 | ···················· | ···················· | ····················· | ···················· | ···················· | ···················· |
| Y | 8003 | hCoV-19/Portugal/PT1646/2020\|EPI_ISL_731921\|2020-1 | ···················· | ···················· | ····················· | ···················· | ···················· | ···················· |
| Y | 8004 | hCoV-19/Portugal/PT1881/2020\|EPI_ISL_731922\|2020-1 | ···················· | ···················· | ····················· | ···················· | ···················· | ···················· |
| Y | 8005 | hCoV-19/Portugal/PT1882/2020\|EPI_ISL_731923\|2020-1 | ···················· | ···················· | ····················· | ···················· | ···················· | ···················· |
| Y | 8006 | hCoV-19/Portugal/PT1914/2020\|EPI_ISL_731924\|2020-1 | ···················· | ···················· | ····················· | ···················· | ···················· | ···················· |
| Y | 8007 | hCoV-19/Portugal/PT1938/2020\|EPI_ISL_731925\|2020-1 | ···················· | ···················· | ····················· | ···················· | ···················· | ···················· |
| Y | 8008 | hCoV-19/Portugal/PT1971/2020\|EPI_ISL_731926\|2020-1 | ···················· | ···················· | ····················· | ···················· | ···················· | ···················· |
| Y | 8009 | hCoV-19/Portugal/PT2012/2020\|EPI_ISL_731927\|2020-1 | ···················· | ···················· | ····················· | ···················· | ···················· | ···················· |
| Y | 8010 | hCoV-19/Portugal/PT2022/2020\|EPI_ISL_731928\|2020-1 | ···················· | ···················· | ····················· | ···················· | ···················· | ···················· |
| Y | 8011 | hCoV-19/Portugal/PT2025/2020\|EPI_ISL_731929\|2020-1 | ···················· | ···················· | ····················· | ···················· | ···················· | ···················· |
| Y | 8012 | hCoV-19/Portugal/PT2026/2020\|EPI_ISL_731930\|2020-1 | ···················· | ···················· | ····················· | ···················· | ···················· | ···················· |
| Y | 8013 | hCoV-19/Portugal/PT1876/2020\|EPI_ISL_731931\|2020-1 | ···················· | ···················· | ····················· | ···················· | ···················· | ···················· |
| Y | 8014 | hCoV-19/Portugal/PT1650/2020\|EPI_ISL_731932\|2020-1 | ···················· | ···················· | ····················· | ···················· | ···················· | ···················· |
| Y | 8015 | hCoV-19/Portugal/PT1677/2020\|EPI_ISL_731933\|2020-1 | ···················· | ···················· | ····················· | ···················· | ···················· | ···················· |
| Y | 8016 | hCoV-19/Portugal/PT1763/2020\|EPI_ISL_731934\|2020-1 | ···················· | ···················· | ····················· | ···················· | ···················· | ···················· |
| Y | 8017 | hCoV-19/Portugal/PT1765/2020\|EPI_ISL_731935\|2020-1 | ···················· | ···················· | ····················· | ···················· | ···················· | ···················· |
| Y | 8018 | hCoV-19/Portugal/PT1790/2020\|EPI_ISL_731936\|2020-1 | ···················· | ···················· | ····················· | ···················· | ···················· | ···················· |
| Y | 8019 | hCoV-19/Portugal/PT1799/2020\|EPI_ISL_731937\|2020-1 | ···················· | ···················· | ····················· | ···················· | ···················· | ···················· |
| Y | 8020 | hCoV-19/Portugal/PT1836/2020\|EPI_ISL_731938\|2020-1 | ···················· | ···················· | ····················· | ···················· | ···················· | ···················· |
| Y | 8021 | hCoV-19/Portugal/PT1896/2020\|EPI_ISL_731939\|2020-1 | ···················· | ···················· | ····················· | ···················· | ···················· | ···················· |
| Y | 8022 | hCoV-19/Portugal/PT1994/2020\|EPI_ISL_731940\|2020-1 | ···················· | ···················· | ····················· | ···················· | ···················· | ···················· |
| Y | 8023 | hCoV-19/Portugal/PT1632/2020\|EPI_ISL_731942\|2020-0 | ···················· | ···················· | ····················· | ···················· | ···················· | ···················· |
| Y | 8024 | hCoV-19/Portugal/PT1633/2020\|EPI_ISL_731943\|2020-0 | ···················· | ···················· | ····················· | ···················· | ···················· | ···················· |
| Y | 8025 | hCoV-19/Portugal/PT1634/2020\|EPI_ISL_731944\|2020-0 | ···················· | ···················· | ····················· | ···················· | ···················· | ···················· |
| Y | 8026 | hCoV-19/Portugal/PT1635/2020\|EPI_ISL_731945\|2020-0 | ···················· | ···················· | ····················· | ···················· | ···················· | ···················· |
| Y | 8027 | hCoV-19/Portugal/PT1636/2020\|EPI_ISL_731946\|2020-0 | ···················· | ···················· | ····················· | ···················· | ···················· | ···················· |
| Y | 8028 | hCoV-19/Portugal/PT1637/2020\|EPI_ISL_731947\|2020-0 | ···················· | ···················· | ····················· | ···················· | ···················· | ···················· |
| Y | 8029 | hCoV-19/Portugal/PT1639/2020\|EPI_ISL_731948\|2020-1 | ···················· | ···················· | ····················· | ···················· | ···················· | ···················· |
| Y | 8030 | hCoV-19/Portugal/PT1640/2020\|EPI_ISL_731949\|2020-1 | ···················· | ···················· | ····················· | ···················· | ···················· | ···················· |
| Y | 8031 | hCoV-19/Portugal/PT1641/2020\|EPI_ISL_731950\|2020-1 | ···················· | ···················· | ····················· | ···················· | ···················· | ···················· |
| Y | 8032 | hCoV-19/Portugal/PT1644/2020\|EPI_ISL_731951\|2020-1 | ···················· | ···················· | ····················· | ···················· | ···················· | ···················· |
| Y | 8033 | hCoV-19/Portugal/PT1647/2020\|EPI_ISL_731952\|2020-1 | ···················· | ···················· | ····················· | ···················· | ···················· | ···················· |
| Y | 8034 | hCoV-19/England/QEUH-C33ADD/2020\|EPI_ISL_734110\|20 | ···················· | ···················· | ····················· | ···················· | ···················· | ···················· |
| Y | 8035 | hCoV-19/England/QEUH-C348C3/2020\|EPI_ISL_734112\|20 | ···················· | ···················· | ····················· | ···················· | ···················· | ···················· |
| Y | 8036 | hCoV-19/England/QEUH-C33C13/2020\|EPI_ISL_734114\|20 | ···················· | ···················· | ····················· | ···················· | ···················· | ···················· |
| Y | 8037 | hCoV-19/England/QEUH-C333B4/2020\|EPI_ISL_734115\|20 | ···················· | ···················· | ····················· | ···················· | ···················· | ···················· |
| Y | 8038 | hCoV-19/England/QEUH-C342A7/2020\|EPI_ISL_734116\|20 | ···················· | ···················· | ····················· | ···················· | ···················· | ···················· |
| Y | 8039 | hCoV-19/England/QEUH-C341B9/2020\|EPI_ISL_734117\|20 | ···················· | ···················· | ····················· | ···················· | ···················· | ···················· |
| Y | 8040 | hCoV-19/England/QEUH-C33A64/2020\|EPI_ISL_734118\|20 | ···················· | ···················· | ····················· | ···················· | ···················· | ···················· |
| Y | 8041 | hCoV-19/England/QEUH-C34122/2020\|EPI_ISL_734120\|20 | ···················· | ···················· | ····················· | ···················· | ···················· | ···················· |
| Y | 8042 | hCoV-19/England/QEUH-C3333C/2020\|EPI_ISL_734121\|20 | ···················· | ···················· | ····················· | ···················· | ···················· | ···················· |
| Y | 8043 | hCoV-19/England/QEUH-C34711/2020\|EPI_ISL_734122\|20 | ···················· | ···················· | ····················· | ···················· | ···················· | ···················· |
| Y | 8044 | hCoV-19/England/QEUH-C3327B/2020\|EPI_ISL_734123\|20 | ···················· | ···················· | ····················· | ···················· | ···················· | ···················· |
| Y | 8045 | hCoV-19/England/QEUH-C333A5/2020\|EPI_ISL_734124\|20 | ···················· | ···················· | ····················· | ···················· | ···················· | ···················· |
| Y | 8046 | hCoV-19/England/QEUH-C3318D/2020\|EPI_ISL_734125\|20 | ···················· | ···················· | ····················· | ···················· | ······T············· | ···················· |
| Y | 8047 | hCoV-19/England/QEUH-C34553/2020\|EPI_ISL_734127\|20 | ···················· | ···················· | ····················· | ···················· | ···················· | ···················· |
| Y | 8048 | hCoV-19/England/QEUH-C34465/2020\|EPI_ISL_734129\|20 | ···················· | ···················· | ····················· | ···················· | ···················· | ···················· |
| Y | 8049 | hCoV-19/England/QEUH-C32EE1/2020\|EPI_ISL_734130\|20 | ···················· | ···················· | ····················· | ···················· | ···················· | ···················· |
| Y | 8050 | hCoV-19/England/QEUH-C341E6/2020\|EPI_ISL_734131\|20 | ···················· | ···················· | ····················· | ···················· | ······T············· | ···················· |
| Y | 8051 | hCoV-19/England/QEUH-C34720/2020\|EPI_ISL_734133\|20 | ···················· | ···················· | ····················· | ···················· | ···················· | ···················· |
| Y | 8052 | hCoV-19/England/QEUH-C33ACE/2020\|EPI_ISL_734134\|20 | ···················· | ···················· | ····················· | ···················· | ···················· | ···················· |
| Y | 8053 | hCoV-19/England/QEUH-C3332D/2020\|EPI_ISL_734135\|20 | ···················· | ···················· | ····················· | ···················· | ···················· | ···················· |
| Y | 8054 | hCoV-19/England/QEUH-C332F3/2020\|EPI_ISL_734136\|20 | ···················· | ···················· | ····················· | ···················· | ···················· | ···················· |
| Y | 8055 | hCoV-19/England/QEUH-C343FF/2020\|EPI_ISL_734137\|20 | ···················· | ···················· | ····················· | ···················· | ···················· | ···················· |
| Y | 8056 | hCoV-19/England/QEUH-C3481E/2020\|EPI_ISL_734138\|20 | ···················· | ···················· | ····················· | ···················· | ···················· | ···················· |
| Y | 8057 | hCoV-19/England/QEUH-C33211/2020\|EPI_ISL_734139\|20 | ···················· | ···················· | ····················· | ···················· | ···················· | ···················· |
| Y | 8058 | hCoV-19/England/QEUH-C3382E/2020\|EPI_ISL_734140\|20 | ···················· | ···················· | ····················· | ···················· | ···················· | ···················· |
| Y | 8059 | hCoV-19/England/QEUH-C33D89/2020\|EPI_ISL_734141\|20 | ···················· | ···················· | ····················· | ···················· | ······T············· | ···················· |

| | | | | | | | |
|---|---|---|---|---|---|---|---|
| Y | 8060 hCoV-19/England/QEUH-C33ABF/2020\|EPI_ISL_734142\|20 | ···················· | ···················· | ···················· | ···················· | ···················· | ···················· |
| Y | 8061 hCoV-19/England/QEUH-C3419B/2020\|EPI_ISL_734145\|20 | ···················· | ···················· | ···················· | ···················· | ···················· | ···················· |
| Y | 8062 hCoV-19/England/QEUH-C338A6/2020\|EPI_ISL_734146\|20 | ···················· | ···················· | ···················· | ···················· | ···················· | ···················· |
| Y | 8063 hCoV-19/England/QEUH-C3379A/2020\|EPI_ISL_734147\|20 | ···················· | ···················· | ···················· | ···················· | ···················· | ···················· |
| Y | 8064 hCoV-19/England/QEUH-C34517/2020\|EPI_ISL_734149\|20 | ···················· | ···················· | ···················· | ···················· | ···················· | ···················· |
| Y | 8065 hCoV-19/England/QEUH-C33DF2/2020\|EPI_ISL_734151\|20 | ···················· | ···················· | ···················· | ···················· | ······T············· | ···················· |
| Y | 8066 hCoV-19/England/QEUH-C32F2A/2020\|EPI_ISL_734152\|20 | ···················· | ···················· | ···················· | ···················· | ···················· | ···················· |
| Y | 8067 hCoV-19/England/QEUH-C33BCB/2020\|EPI_ISL_734154\|20 | ···················· | ···················· | ···················· | ···················· | ···················· | ···················· |
| Y | 8068 hCoV-19/England/QEUH-C34878/2020\|EPI_ISL_734155\|20 | ···················· | ···················· | ···················· | ···················· | ···················· | ···················· |
| Y | 8069 hCoV-19/England/QEUH-C33E77/2020\|EPI_ISL_734156\|20 | ···················· | ···················· | ···················· | ···················· | ···················· | ···················· |
| Y | 8070 hCoV-19/England/QEUH-C33730/2020\|EPI_ISL_734157\|20 | ···················· | ···················· | ···················· | ···················· | ···················· | ···················· |
| Y | 8071 hCoV-19/England/QEUH-C34025/2020\|EPI_ISL_734158\|20 | ···················· | ···················· | ···················· | ···················· | ···················· | ···················· |
| Y | 8072 hCoV-19/England/QEUH-C33B61/2020\|EPI_ISL_734159\|20 | ···················· | ···················· | ···················· | ···················· | ···················· | ···················· |
| Y | 8073 hCoV-19/England/QEUH-C33CE6/2020\|EPI_ISL_734161\|20 | ···················· | ·A·······A·········· | ···················· | ···················· | ···················· | ···················· |
| Y | 8074 hCoV-19/England/QEUH-C34580/2020\|EPI_ISL_734162\|20 | ···················· | ···················· | ···················· | ···················· | ···················· | ···················· |
| Y | 8075 hCoV-19/France/BRE-IPP9985/2020\|EPI_ISL_734168\|202 | ···················· | ···················· | ···················· | ···················· | ···················· | ···················· |
| Y | 8076 hCoV-19/France/BRE-IPP9997/2020\|EPI_ISL_734169\|202 | ···················· | ···················· | ···················· | ···················· | ···················· | ···················· |
| Y | 8077 hCoV-19/USA/VA-DCLS-2399/2020\|EPI_ISL_734170\|2020- | ···················· | ···················· | ···················· | ···················· | ·········T·········· | ···················· |
| Y | 8078 hCoV-19/USA/VA-DCLS-2400/2020\|EPI_ISL_734171\|2020- | ···················· | ···················· | ···················· | ···················· | ·········T·········· | ···················· |
| Y | 8079 hCoV-19/USA/VA-DCLS-2401/2020\|EPI_ISL_734172\|2020- | ···················· | ···················· | ···················· | ···················· | ·········T·········· | ···················· |
| Y | 8080 hCoV-19/USA/VA-DCLS-2402/2020\|EPI_ISL_734173\|2020- | ···················· | ···················· | ···················· | ···················· | ·········T·········· | ···················· |
| Y | 8081 hCoV-19/USA/VA-DCLS-2403/2020\|EPI_ISL_734174\|2020- | ···················· | ···················· | ···················· | ···················· | ·········T·········· | ···················· |
| Y | 8082 hCoV-19/USA/VA-DCLS-2404/2020\|EPI_ISL_734175\|2020- | ···················· | ···················· | ···················· | ···················· | ·········T·········· | ···················· |
| Y | 8083 hCoV-19/USA/VA-DCLS-2405/2020\|EPI_ISL_734176\|2020- | ···················· | ···················· | ···················· | ···················· | ·········T·········· | ···················· |
| Y | 8084 hCoV-19/USA/VA-DCLS-2406/2020\|EPI_ISL_734177\|2020- | ···················· | ···················· | ···················· | ···················· | ·········T·········· | ···················· |
| Y | 8085 hCoV-19/USA/VA-DCLS-2407/2020\|EPI_ISL_734178\|2020- | ···················· | ···················· | ···················· | ···················· | ·········T·········· | ···················· |
| Y | 8086 hCoV-19/USA/VA-DCLS-2408/2020\|EPI_ISL_734179\|2020- | ···················· | ···················· | ···················· | ···················· | ·········T·········· | ···················· |
| Y | 8087 hCoV-19/USA/VA-DCLS-2409/2020\|EPI_ISL_734180\|2020- | ···················· | ···················· | ···················· | ···················· | ···················· | ···················· |
| Y | 8088 hCoV-19/USA/VA-DCLS-2410/2020\|EPI_ISL_734181\|2020- | ···················· | ···················· | ···················· | ···················· | ···················· | ···················· |
| Y | 8089 hCoV-19/USA/VA-DCLS-2411/2020\|EPI_ISL_734182\|2020- | ···················· | ···················· | ···················· | ···················· | ···················· | ···················· |
| Y | 8090 hCoV-19/USA/VA-DCLS-2412/2020\|EPI_ISL_734183\|2020- | ···················· | ···················· | ···················· | ···················· | ···················· | ···················· |
| Y | 8091 hCoV-19/USA/VA-DCLS-2413/2020\|EPI_ISL_734184\|2020- | ···················· | ···················· | ···················· | ···················· | ···················· | ···················· |
| Y | 8092 hCoV-19/USA/VA-DCLS-2434/2020\|EPI_ISL_734185\|2020- | ···················· | ···················· | ···················· | ···················· | ···················· | ···················· |
| Y | 8093 hCoV-19/USA/VA-DCLS-2437/2020\|EPI_ISL_734186\|2020- | ···················· | ······A············· | ···················· | ···················· | ···················· | ···················· |
| Y | 8094 hCoV-19/USA/VA-DCLS-2438/2020\|EPI_ISL_734187\|2020- | ···················· | ······A············· | ···················· | ···················· | ···················· | ···················· |
| Y | 8095 hCoV-19/USA/VA-DCLS-2439/2020\|EPI_ISL_734188\|2020- | ···················· | ······A············· | ···················· | ···················· | ···················· | ···················· |
| Y | 8096 hCoV-19/USA/VA-DCLS-2440/2020\|EPI_ISL_734189\|2020- | ···················· | ······A············· | ···················· | ···················· | ···················· | ···················· |
| Y | 8097 hCoV-19/USA/VA-DCLS-2441/2020\|EPI_ISL_734190\|2020- | ···················· | ······A············· | ···················· | ···················· | ···················· | ···················· |
| Y | 8098 hCoV-19/USA/VA-DCLS-2442/2020\|EPI_ISL_734191\|2020- | ···················· | ···················· | ···················· | ···················· | ···················· | ···················· |
| Y | 8099 hCoV-19/Brazil/SP-739/2020\|EPI_ISL_735400\|2020-07- | ···················· | ···················· | ···················· | ···················· | ···················· | ···················· |
| Y | 8100 hCoV-19/Brazil/SP-740/2020\|EPI_ISL_735401\|2020-07- | ···················· | ···················· | ···················· | ···················· | ···················· | ···················· |
| Y | 8101 hCoV-19/Brazil/SP-742/2020\|EPI_ISL_735402\|2020-07- | ···················· | ···················· | ···················· | ···················· | ···················· | ···················· |
| Y | 8102 hCoV-19/Brazil/SP-743/2020\|EPI_ISL_735403\|2020-07- | ···················· | ···················· | ···················· | ···················· | ···················· | ···················· |
| Y | 8103 hCoV-19/Brazil/SP-744/2020\|EPI_ISL_735404\|2020-07- | ···················· | ···················· | ···················· | ···················· | ···················· | ···················· |
| Y | 8104 hCoV-19/Brazil/SP-745/2020\|EPI_ISL_735405\|2020-06- | ···················· | ···················· | ···················· | ···················· | ···················· | ···················· |
| Y | 8105 hCoV-19/Brazil/SP-748/2020\|EPI_ISL_735406\|2020-07- | ···················· | ···················· | ···················· | ···················· | ···················· | ···················· |
| Y | 8106 hCoV-19/Brazil/SP-749/2020\|EPI_ISL_735407\|2020-06- | ···················· | ···················· | ···········T········ | ···················· | ···················· | ···················· |
| Y | 8107 hCoV-19/Brazil/SP-750/2020\|EPI_ISL_735408\|2020-06- | ···················· | ···················· | ···················· | ···················· | ···················· | ···················· |
| Y | 8108 hCoV-19/Brazil/SP-751/2020\|EPI_ISL_735409\|2020-06- | ···················· | ···················· | ···················· | ···················· | ···················· | ···················· |
| Y | 8109 hCoV-19/Brazil/SP-754/2020\|EPI_ISL_735410\|2020-07- | ···················· | ···················· | ···················· | ···················· | ···················· | ···················· |
| Y | 8110 hCoV-19/Brazil/SP-756/2020\|EPI_ISL_735411\|2020-07- | ···················· | ···················· | ···················· | ···················· | ···················· | ···················· |
| Y | 8111 hCoV-19/Brazil/SP-758/2020\|EPI_ISL_735412\|2020-06- | ···················· | ···················· | ···················· | ···················· | ···················· | ···················· |
| Y | 8112 hCoV-19/Brazil/SP-759/2020\|EPI_ISL_735413\|2020-06- | ···················· | ···················· | ···················· | ···················· | ···················· | ···················· |
| Y | 8113 hCoV-19/Brazil/SP-761/2020\|EPI_ISL_735414\|2020-06- | ···················· | ···················· | ···················· | ···················· | ···················· | ···················· |
| Y | 8114 hCoV-19/Brazil/SP-762/2020\|EPI_ISL_735415\|2020-06- | ···················· | ···················· | ···················· | ···················· | ···················· | ···················· |
| Y | 8115 hCoV-19/Brazil/SP-764/2020\|EPI_ISL_735416\|2020-06- | ···················· | ···················· | ···················· | ···················· | ···················· | ···················· |
| Y | 8116 hCoV-19/Brazil/SP-765/2020\|EPI_ISL_735417\|2020-06- | ···················· | ···················· | ···················· | ···················· | ···················· | ···················· |
| Y | 8117 hCoV-19/Brazil/SP-766/2020\|EPI_ISL_735418\|2020-06- | ···················· | ···················· | ···················· | ···················· | ···················· | ···················· |
| Y | 8118 hCoV-19/Brazil/SP-770/2020\|EPI_ISL_735419\|2020-07- | ···················· | ···················· | ···················· | ···················· | ···················· | ···················· |
| Y | 8119 hCoV-19/Brazil/SP-772/2020\|EPI_ISL_735420\|2020-07- | ···················· | ···················· | ···················· | ···················· | ···················· | ···················· |
| Y | 8120 hCoV-19/Brazil/SP-773/2020\|EPI_ISL_735421\|2020-07- | ···················· | ···················· | ···················· | ···················· | ···················· | ···················· |
| Y | 8121 hCoV-19/Brazil/SP-774/2020\|EPI_ISL_735422\|2020-06- | ···················· | ···················· | ···················· | ···················· | ···················· | ···················· |
| Y | 8122 hCoV-19/Brazil/SP-783/2020\|EPI_ISL_735423\|2020-07- | ···················· | ···················· | ···················· | ···················· | ···················· | ···················· |
| Y | 8123 hCoV-19/Brazil/SP-784/2020\|EPI_ISL_735424\|2020-07- | ···················· | ···················· | ···················· | ···················· | ···················· | ···················· |
| Y | 8124 hCoV-19/Brazil/SP-785/2020\|EPI_ISL_735425\|2020-06- | ···················· | ···················· | ···················· | ···················· | ···················· | ···················· |
| Y | 8125 hCoV-19/Brazil/SP-786/2020\|EPI_ISL_735426\|2020-07- | ···················· | ···················· | ···················· | ···················· | ···················· | ···················· |
| Y | 8126 hCoV-19/Brazil/SP-789/2020\|EPI_ISL_735427\|2020-07- | ···················· | ···················· | ···················· | ···················· | ···················· | ···················· |
| Y | 8127 hCoV-19/Brazil/SP-791/2020\|EPI_ISL_735428\|2020-07- | ···················· | ···················· | ···················· | ···················· | ···················· | ···················· |
| Y | 8128 hCoV-19/Rwanda/NRLNAT1004/2020\|EPI_ISL_735444\|2020 | ···················· | ·A·················· | ···················· | ···················· | ···················· | ···················· |
| Y | 8129 hCoV-19/Rwanda/NRLNAT1008/2020\|EPI_ISL_735448\|2020 | ···················· | ···················· | ···················· | ···················· | ···················· | ···················· |
| Y | 8130 hCoV-19/USA/WA-UW-1492/2020\|EPI_ISL_735449\|2020-03 | ···················· | ···················· | ···················· | ···················· | ···················· | ···················· |
| Y | 8131 hCoV-19/USA/WA-UW-1967/2020\|EPI_ISL_735450\|2020-03 | ···················· | ···················· | ···················· | ···················· | ···················· | ···················· |
| Y | 8132 hCoV-19/USA/WA-UW-29724/2020\|EPI_ISL_735451\|2020-0 | ···················· | ···················· | ···················· | ···················· | ···················· | ···················· |
| Y | 8133 hCoV-19/USA/WA-UW-30834/2020\|EPI_ISL_735452\|2020-1 | ···················· | ···················· | ···················· | ···················· | ···················· | ···················· |
| Y | 8134 hCoV-19/USA/WA-UW-30835/2020\|EPI_ISL_735453\|2020-1 | ···················· | ·A·················· | ···················· | ···················· | ···················· | ···················· |
| Y | 8135 hCoV-19/USA/WA-UW-30922/2020\|EPI_ISL_735457\|2020-1 | ···················· | ·A·················· | ···················· | ···················· | ···················· | ···················· |
| Y | 8136 hCoV-19/USA/WA-UW-30946/2020\|EPI_ISL_735458\|2020-1 | ···················· | ·A·················· | ···················· | ···················· | ···················· | ···················· |
| Y | 8137 hCoV-19/USA/WA-UW-31203/2020\|EPI_ISL_735459\|2020-1 | ···················· | ···················· | ···················· | ···················· | ···················· | ···················· |
| Y | 8138 hCoV-19/USA/WA-UW-31214/2020\|EPI_ISL_735461\|2020-1 | ···················· | ·A·················· | ···················· | ···················· | ···················· | ···················· |
| Y | 8139 hCoV-19/USA/WA-UW-31218/2020\|EPI_ISL_735462\|2020-1 | ···················· | ·A·················· | ···················· | ···················· | ···················· | ···················· |

| | | | | | | | |
|---|---|---|---|---|---|---|---|
| Y | 8140 hCoV-19/USA/WA-UW-31219/2020\|EPI_ISL_735463\|2020-1 | ···················· | ···················· | ···················· | ···················· | ···················· | ···················· |
| Y | 8142 hCoV-19/USA/WA-UW-31224/2020\|EPI_ISL_735465\|2020-1 | ···················· | ·A·················· | ···················· | ···················· | ···················· | ···················· |
| Y | 8143 hCoV-19/USA/UNKNOWN-UW-31229/2020\|EPI_ISL_735466\|2 | ···················· | ···················· | ···················· | ···················· | ···················· | ···················· |
| Y | 8144 hCoV-19/USA/WA-UW-31239/2020\|EPI_ISL_735469\|2020-1 | ···················· | ·A·················· | ···················· | ···················· | ···················· | ···················· |
| Y | 8145 hCoV-19/USA/WA-UW-31251/2020\|EPI_ISL_735470\|2020-1 | ···················· | ···················· | ···················· | ···················· | ···················· | ···················· |
| Y | 8146 hCoV-19/USA/WA-UW-31382/2020\|EPI_ISL_735472\|2020-1 | ···················· | ···················· | ···················· | ···················· | ···················· | ···················· |
| Y | 8147 hCoV-19/USA/WA-UW-31402/2020\|EPI_ISL_735473\|2020-1 | ···················· | ···················· | ···················· | ···················· | ···················· | ···················· |
| Y | 8148 hCoV-19/USA/WA-UW-31404/2020\|EPI_ISL_735474\|2020-1 | ···················· | ···················· | ···················· | ···················· | ···················· | ···················· |
| Y | 8149 hCoV-19/USA/WA-UW-31407/2020\|EPI_ISL_735475\|2020-1 | ···················· | ···················· | ···················· | ···················· | ···················· | ···················· |
| Y | 8150 hCoV-19/USA/WA-UW-31410/2020\|EPI_ISL_735477\|2020-1 | ···················· | ···················· | ···················· | ···················· | ···················· | ···················· |
| Y | 8151 hCoV-19/USA/WA-UW-31411/2020\|EPI_ISL_735478\|2020-1 | ···················· | ·A·················· | ···················· | ···················· | ···················· | ···················· |
| Y | 8152 hCoV-19/USA/WA-UW-31416/2020\|EPI_ISL_735479\|2020-1 | ···················· | ···················· | ···················· | ···················· | ···················· | ···················· |
| Y | 8153 hCoV-19/USA/WA-UW-31428/2020\|EPI_ISL_735482\|2020-1 | ···················· | ···················· | ···················· | ···················· | ···················· | ···················· |
| Y | 8154 hCoV-19/USA/WA-UW-31441/2020\|EPI_ISL_735484\|2020-1 | ···················· | ···················· | ···················· | ···················· | ···················· | ···················· |
| Y | 8155 hCoV-19/USA/WA-UW-31591/2020\|EPI_ISL_735485\|2020-1 | ···················· | ·A·················· | ···················· | ···················· | ···················· | ···················· |
| Y | 8156 hCoV-19/USA/WA-UW-31637/2020\|EPI_ISL_735489\|2020-1 | ···················· | ···················· | ···················· | ···················· | ···················· | ···················· |
| Y | 8157 hCoV-19/Bangladesh/CU-CTG-02/2020\|EPI_ISL_735490\|2 | ···················· | ···················· | ···················· | ···················· | ···················· | ···················· |
| Y | 8158 hCoV-19/Bangladesh/CU-CTG-04/2020\|EPI_ISL_735492\|2 | ···················· | ···················· | ···················· | ···················· | ···················· | ···················· |
| Y | 8159 hCoV-19/Bangladesh/CU-CTG-09/2020\|EPI_ISL_735493\|2 | ···················· | ···················· | ···················· | ···················· | ···················· | ···················· |
| Y | 8160 hCoV-19/Bangladesh/CU-CTG-13/2020\|EPI_ISL_735494\|2 | ···················· | ···················· | ···················· | ···················· | ···················· | ···················· |
| Y | 8161 hCoV-19/Bangladesh/CU-CTG-18/2020\|EPI_ISL_735495\|2 | ···················· | ···················· | ···················· | ···················· | ···················· | ···················· |
| Y | 8162 hCoV-19/Bangladesh/CU-CTG-24/2020\|EPI_ISL_735496\|2 | ···················· | ·A·················· | ···················· | ···················· | ···················· | ···A················ |
| Y | 8163 hCoV-19/Bangladesh/CU-CTG-32/2020\|EPI_ISL_735497\|2 | ···················· | ···················· | ···················· | ···················· | ···················· | ···················· |
| Y | 8164 hCoV-19/Bangladesh/CU-CTG-38/2020\|EPI_ISL_735498\|2 | ···················· | ···················· | ···················· | ···················· | ···················· | ···················· |
| Y | 8165 hCoV-19/Bangladesh/CU-CTG-44/2020\|EPI_ISL_735499\|2 | ···················· | ···················· | ···················· | ···················· | ···················· | ···················· |
| Y | 8166 hCoV-19/England/ALDP-C48A7B/2020\|EPI_ISL_736729\|20 | ···················· | ···················· | ···················· | ···················· | ···················· | ···················· |
| Y | 8167 hCoV-19/England/ALDP-C483BC/2020\|EPI_ISL_736741\|20 | ···················· | ···················· | ···················· | ···················· | ···················· | ···················· |
| Y | 8168 hCoV-19/England/MILK-C508D0/2020\|EPI_ISL_736744\|20 | ···················· | ···················· | ···················· | ···················· | ···················· | ···················· |
| Y | 8169 hCoV-19/England/QEUH-C4BD24/2020\|EPI_ISL_736745\|20 | ···················· | ···················· | ···················· | ···················· | ···················· | ···················· |
| Y | 8170 hCoV-19/England/ALDP-C48BB4/2020\|EPI_ISL_736748\|20 | ···················· | ···················· | ···················· | ···················· | ···················· | ···················· |
| Y | 8171 hCoV-19/England/ALDP-C48835/2020\|EPI_ISL_736751\|20 | ···················· | ···················· | ···················· | ···················· | ···················· | ···················· |
| Y | 8172 hCoV-19/England/QEUH-C4BD8E/2020\|EPI_ISL_736752\|20 | ···················· | ···················· | ···················· | ···················· | ···················· | ···················· |
| Y | 8173 hCoV-19/England/QEUH-C4BB48/2020\|EPI_ISL_736754\|20 | ···················· | ···················· | ···················· | ···················· | ···················· | ···················· |
| Y | 8174 hCoV-19/England/ALDP-C48BC3/2020\|EPI_ISL_736755\|20 | ···················· | ···················· | ···················· | ···················· | ···················· | ···················· |
| Y | 8175 hCoV-19/England/ALDP-C48413/2020\|EPI_ISL_736756\|20 | ···················· | ···················· | ···················· | ···················· | ······T············· | ···················· |
| Y | 8176 hCoV-19/England/ALDP-C3A9F7/2020\|EPI_ISL_736758\|20 | ···················· | ···················· | ···················· | ···················· | ···················· | ···················· |
| Y | 8177 hCoV-19/England/ALDP-C3D129/2020\|EPI_ISL_736770\|20 | ···················· | ···················· | ···················· | ···················· | ···················· | ···················· |
| Y | 8178 hCoV-19/England/ALDP-C3D2BD/2020\|EPI_ISL_736771\|20 | ···················· | ···················· | ···················· | ···················· | ···················· | ···················· |
| Y | 8179 hCoV-19/England/ALDP-C24486/2020\|EPI_ISL_736772\|20 | ···················· | ···················· | ···················· | ···················· | ···················· | ···················· |
| Y | 8180 hCoV-19/England/QEUH-C336D9/2020\|EPI_ISL_736774\|20 | ···················· | ···················· | ···················· | ···················· | ···················· | ···················· |
| Y | 8181 hCoV-19/USA/WA-UW-1483/2020\|EPI_ISL_736775\|2020-03 | ···················· | ···················· | ···················· | ···················· | ···················· | ···················· |
| Y | 8182 hCoV-19/USA/WA-UW-30840/2020\|EPI_ISL_736776\|2020-1 | ···················· | ···················· | ···················· | ···················· | ···················· | ···················· |
| Y | 8183 hCoV-19/Italy/CAM-TIGEM-55/2020\|EPI_ISL_736789\|202 | ···················· | ···················· | ···················· | ···················· | ······T············· | ···················· |
| Y | 8184 hCoV-19/Italy/CAM-TIGEM-1/2020\|EPI_ISL_736791\|2020 | ···················· | ···················· | ···················· | ···················· | ···················· | ···················· |
| Y | 8185 hCoV-19/Italy/CAM-TIGEM-2/2020\|EPI_ISL_736792\|2020 | ···················· | ···················· | ···················· | ···················· | ···················· | ···················· |
| Y | 8186 hCoV-19/Italy/CAM-TIGEM-3/2020\|EPI_ISL_736793\|2020 | ···················· | ·A·················· | ···················· | ···················· | ···················· | ···················· |
| Y | 8187 hCoV-19/Italy/CAM-TIGEM-4/2020\|EPI_ISL_736794\|2020 | ···················· | ·A·················· | ···················· | ···················· | ···················· | ···················· |
| Y | 8188 hCoV-19/Italy/CAM-TIGEM-5/2020\|EPI_ISL_736795\|2020 | ···················· | ·A·················· | ···················· | ···················· | ···················· | ···················· |
| Y | 8189 hCoV-19/Italy/CAM-TIGEM-7/2020\|EPI_ISL_736796\|2020 | ···················· | ···················· | ···················· | ···················· | ···················· | ···················· |
| Y | 8190 hCoV-19/Italy/CAM-TIGEM-8/2020\|EPI_ISL_736797\|2020 | ···················· | ···················· | ···················· | ···················· | ···················· | ···················· |
| Y | 8191 hCoV-19/Italy/CAM-TIGEM-9/2020\|EPI_ISL_736798\|2020 | ···················· | ···················· | ···················· | ···················· | ···················· | ···················· |
| Y | 8192 hCoV-19/Italy/CAM-TIGEM-11/2020\|EPI_ISL_736799\|202 | ···················· | ···················· | ···················· | ···················· | ···················· | ···················· |
| Y | 8193 hCoV-19/Wales/PHWC-497F49/2020\|EPI_ISL_727000\|2020 | ···················· | ···················· | ···················· | ···················· | ···················· | ···················· |
| Y | 8194 hCoV-19/Wales/PHWC-497F58/2020\|EPI_ISL_727001\|2020 | ···················· | ···················· | ···················· | ···················· | ···················· | ···················· |
| Y | 8195 hCoV-19/Wales/PHWC-497F67/2020\|EPI_ISL_727002\|2020 | ···················· | ···················· | ···················· | ···················· | ···················· | ···················· |
| Y | 8196 hCoV-19/Wales/PHWC-497F85/2020\|EPI_ISL_727004\|2020 | ···················· | ···················· | ···················· | ···················· | ···················· | ···················· |
| Y | 8197 hCoV-19/Wales/PHWC-497F94/2020\|EPI_ISL_727005\|2020 | ···················· | ···················· | ···················· | ···················· | ···················· | ···················· |
| Y | 8198 hCoV-19/Wales/PHWC-497FA3/2020\|EPI_ISL_727006\|2020 | ···················· | ···················· | ···················· | ···················· | ···················· | ···················· |
| Y | 8200 hCoV-19/Wales/PHWC-497FC1/2020\|EPI_ISL_727008\|2020 | ···················· | ···················· | ···················· | ···················· | ···················· | ···················· |
| Y | 8201 hCoV-19/Wales/PHWC-497FD0/2020\|EPI_ISL_727009\|2020 | ···················· | ···················· | ···················· | ···················· | ···················· | ···················· |
| Y | 8203 hCoV-19/Wales/PHWC-497FFE/2020\|EPI_ISL_727011\|2020 | ···················· | ···················· | ···················· | ···················· | ···················· | ···················· |
| Y | 8204 hCoV-19/Wales/PHWC-498027/2020\|EPI_ISL_727013\|2020 | ···················· | ······A············· | ···················· | ···················· | ···················· | ···················· |
| Y | 8205 hCoV-19/Wales/PHWC-498036/2020\|EPI_ISL_727014\|2020 | ···················· | ···················· | ···················· | ···················· | ···················· | ···················· |
| Y | 8206 hCoV-19/Wales/PHWC-498045/2020\|EPI_ISL_727015\|2020 | ···················· | ···················· | ···················· | ···················· | ···················· | ···················· |
| Y | 8207 hCoV-19/Wales/PHWC-498054/2020\|EPI_ISL_727016\|2020 | ···················· | ···················· | ···················· | ···················· | ···················· | ···················· |
| Y | 8208 hCoV-19/Wales/PHWC-498063/2020\|EPI_ISL_727017\|2020 | ···················· | ···················· | ···················· | ···················· | ···················· | ···················· |
| Y | 8209 hCoV-19/Wales/PHWC-498072/2020\|EPI_ISL_727018\|2020 | ···················· | ···················· | ···················· | ···················· | ···················· | ···················· |
| Y | 8210 hCoV-19/Wales/PHWC-498081/2020\|EPI_ISL_727019\|2020 | ···················· | ···················· | ···················· | ···················· | ···················· | ···················· |
| Y | 8211 hCoV-19/Wales/PHWC-498090/2020\|EPI_ISL_727020\|2020 | ···················· | ···················· | ···················· | ···················· | ···················· | ···················· |
| Y | 8212 hCoV-19/Wales/PHWC-4980AF/2020\|EPI_ISL_727021\|2020 | ···················· | ···················· | ···················· | ···················· | ···················· | ···················· |
| Y | 8213 hCoV-19/Wales/PHWC-4980CD/2020\|EPI_ISL_727023\|2020 | ···················· | ···················· | ···················· | ···················· | ···················· | ···················· |
| Y | 8214 hCoV-19/Wales/PHWC-4980DC/2020\|EPI_ISL_727024\|2020 | ···················· | ···················· | ···················· | ···················· | ···················· | ···················· |
| Y | 8215 hCoV-19/Wales/PHWC-4980EB/2020\|EPI_ISL_727025\|2020 | ···················· | ···················· | ···················· | ···················· | ···················· | ···················· |
| Y | 8216 hCoV-19/Wales/PHWC-4980FA/2020\|EPI_ISL_727026\|2020 | ···················· | ···················· | ···················· | ···················· | ···················· | ···················· |
| Y | 8217 hCoV-19/Wales/PHWC-498106/2020\|EPI_ISL_727027\|2020 | ···················· | ···················· | ···················· | ···················· | ···················· | ···················· |
| Y | 8219 hCoV-19/Wales/PHWC-498124/2020\|EPI_ISL_727029\|2020 | ···················· | ···················· | ···················· | ···················· | ···················· | ···················· |
| Y | 8220 hCoV-19/Wales/PHWC-498203/2020\|EPI_ISL_727040\|2020 | ···················· | ···················· | ···················· | ···················· | ···················· | ···················· |
| Y | 8221 hCoV-19/Wales/PHWC-498212/2020\|EPI_ISL_727041\|2020 | ···················· | ···················· | ···················· | ···················· | ···················· | ···················· |
| Y | 8222 hCoV-19/Wales/PHWC-498221/2020\|EPI_ISL_727042\|2020 | ···················· | ···················· | ···················· | ···················· | ···················· | ···················· |
| Y | 8223 hCoV-19/Wales/PHWC-49824F/2020\|EPI_ISL_727044\|2020 | ···················· | ···················· | ···················· | ···················· | ···················· | ···················· |

| | | | | | | | | |
|---|---|---|---|---|---|---|---|---|
| Y | 8224 | hCoV-19/Wales/PHWC-49826D/2020\|EPI_ISL_727045\|2020 | ···················· | ···················· | ···················· | ···················· | ···················· | ···················· |
| Y | 8225 | hCoV-19/Wales/PHWC-49827C/2020\|EPI_ISL_727046\|2020 | ···················· | ···················· | ···················· | ···················· | ···················· | ···················· |
| Y | 8226 | hCoV-19/Wales/PHWC-49828B/2020\|EPI_ISL_727047\|2020 | ···················· | ···················· | ···················· | ···················· | ···················· | ···················· |
| Y | 8227 | hCoV-19/Wales/PHWC-4982A9/2020\|EPI_ISL_727049\|2020 | ···················· | ···················· | ···················· | ···················· | ···················· | ···················· |
| Y | 8228 | hCoV-19/Wales/PHWC-4982B8/2020\|EPI_ISL_727050\|2020 | ···················· | ···················· | ···················· | ···················· | ···················· | ···················· |
| Y | 8229 | hCoV-19/Wales/PHWC-4982C7/2020\|EPI_ISL_727051\|2020 | ···················· | ···················· | ···················· | ···················· | ···················· | ···················· |
| Y | 8230 | hCoV-19/Wales/PHWC-4982E5/2020\|EPI_ISL_727053\|2020 | ···················· | ···················· | ···················· | ···················· | ···················· | ···················· |
| Y | 8231 | hCoV-19/Wales/PHWC-4982F4/2020\|EPI_ISL_727054\|2020 | ···················· | ···················· | ···················· | ···················· | ···················· | ···················· |
| Y | 8232 | hCoV-19/Wales/PHWC-498300/2020\|EPI_ISL_727055\|2020 | ···················· | ···················· | ···················· | ···················· | ···················· | ···················· |
| Y | 8233 | hCoV-19/Wales/PHWC-49831F/2020\|EPI_ISL_727056\|2020 | ···················· | ······A············· | ···················· | ···················· | ···················· | ···················· |
| Y | 8234 | hCoV-19/Wales/PHWC-49832E/2020\|EPI_ISL_727057\|2020 | ···················· | ···················· | ···················· | ···················· | ···················· | ···················· |
| Y | 8235 | hCoV-19/Wales/PHWC-49833D/2020\|EPI_ISL_727058\|2020 | ···················· | ···················· | ···················· | ···················· | ···················· | ···················· |
| Y | 8236 | hCoV-19/Wales/PHWC-49834C/2020\|EPI_ISL_727059\|2020 | ···················· | ···················· | ···················· | ···················· | ···················· | ···················· |
| Y | 8237 | hCoV-19/France/ARA-107339/2020\|EPI_ISL_728310\|2020 | ···················· | ···················· | ···················· | ···················· | ···················· | ···················· |
| Y | 8238 | hCoV-19/France/ARA-107421/2020\|EPI_ISL_728311\|2020 | ···················· | ···················· | ···················· | ···················· | ···················· | ···················· |
| Y | 8239 | hCoV-19/France/ARA-107506/2020\|EPI_ISL_728312\|2020 | ···················· | ···················· | ···················· | ···················· | ···················· | ···················· |
| Y | 8240 | hCoV-19/France/ARA-107560/2020\|EPI_ISL_728313\|2020 | ···················· | ·A·················· | ···················· | ···················· | ···················· | ···················· |
| Y | 8241 | hCoV-19/France/ARA-107566/2020\|EPI_ISL_728314\|2020 | ···················· | ···················· | ···················· | ···················· | ···················· | ···················· |
| Y | 8242 | hCoV-19/France/ARA-107604/2020\|EPI_ISL_728315\|2020 | ···················· | ···················· | ···················· | ···················· | ···················· | ···················· |
| Y | 8243 | hCoV-19/France/ARA-107712/2020\|EPI_ISL_728316\|2020 | ···················· | ···················· | ···················· | ···················· | ···················· | ···················· |
| Y | 8244 | hCoV-19/France/ARA-107994/2020\|EPI_ISL_728317\|2020 | ···················· | ···················· | ···················· | ···················· | ···················· | ···················· |
| Y | 8245 | hCoV-19/France/ARA-108014/2020\|EPI_ISL_728318\|2020 | ···················· | ···················· | ···················· | ···················· | ···················· | ···················· |
| Y | 8246 | hCoV-19/France/ARA-108039/2020\|EPI_ISL_728319\|2020 | ···················· | ···················· | ···················· | ···················· | ···················· | ···················· |
| Y | 8247 | hCoV-19/France/ARA-108102/2020\|EPI_ISL_728320\|2020 | ···················· | ···················· | ···················· | ···················· | ···················· | ···················· |
| Y | 8248 | hCoV-19/Morocco/FMP-15/2020\|EPI_ISL_728322\|2020-11 | ···················· | ···················· | ···················· | ···················· | ···················· | ···················· |
| Y | 8249 | hCoV-19/Germany/TH-IIMK-CaSe-11/2020\|EPI_ISL_72832 | ···················· | ···················· | ···················· | ···················· | ···················· | ···················· |
| Y | 8250 | hCoV-19/Germany/TH-IIMK-CaSe-12/2020\|EPI_ISL_72832 | ···················· | ···················· | ···················· | ···················· | ···················· | ···················· |
| Y | 8251 | hCoV-19/India/MH-BJMC-8674/2020\|EPI_ISL_728325\|202 | ···················· | ···················· | ···················· | ···················· | ···················· | ···················· |
| Y | 8252 | hCoV-19/India/MH-BJMC-29654/2020\|EPI_ISL_728326\|20 | ···················· | ···················· | ···················· | ···················· | ···················· | ···················· |
| Y | 8253 | hCoV-19/India/MH-BJMC-21565/2020\|EPI_ISL_728327\|20 | ···················· | ···················· | ···················· | ···················· | ···················· | ···················· |
| Y | 8254 | hCoV-19/India/MH-BJMC-8827/2020\|EPI_ISL_728328\|202 | ···················· | ···················· | ···················· | ···················· | ···················· | ···················· |
| Y | 8255 | hCoV-19/Germany/TH-IIMK-CaSe-13/2020\|EPI_ISL_72833 | ···················· | ···················· | ···················· | ···················· | ···················· | ···················· |
| Y | 8256 | hCoV-19/Morocco/FMP-16/2020\|EPI_ISL_728332\|2020-11 | ···················· | ···················· | ···················· | ···················· | ···················· | ···················· |
| Y | 8257 | hCoV-19/Germany/TH-IIMK-CaSe-14/2020\|EPI_ISL_72833 | ···················· | ···················· | ···················· | ···················· | ···················· | ···················· |
| Y | 8258 | hCoV-19/Morocco/FMP-17/2020\|EPI_ISL_728334\|2020-12 | ···················· | ···················· | ···················· | ···················· | ···················· | ···················· |
| Y | 8259 | hCoV-19/Germany/TH-IIMK-CaSe-15/2020\|EPI_ISL_72833 | ···················· | ···················· | ···················· | ···················· | ···················· | ···················· |
| Y | 8260 | hCoV-19/Germany/TH-IIMK-CaSe-16/2020\|EPI_ISL_72833 | ···················· | ···················· | ···················· | ···················· | ···················· | ···················· |
| Y | 8261 | hCoV-19/Germany/TH-IIMK-CaSe-17/2020\|EPI_ISL_72833 | ···················· | ···················· | ···················· | ···················· | ···················· | ···················· |
| Y | 8262 | hCoV-19/Morocco/FMP-19/2020\|EPI_ISL_728339\|2020-12 | ···················· | ···················· | ···················· | ···················· | ···················· | ···················· |
| Y | 8263 | hCoV-19/Morocco/FMP-20/2020\|EPI_ISL_728340\|2020-12 | ···················· | ···················· | ···················· | ···················· | ···················· | ···················· |
| Y | 8264 | hCoV-19/Germany/TH-IIMK-CaSe-18/2020\|EPI_ISL_72834 | ···················· | ···················· | ···················· | ···················· | ···················· | ···················· |
| Y | 8265 | hCoV-19/Morocco/FMP-21/2020\|EPI_ISL_728342\|2020-12 | ···················· | ···················· | ···················· | ···················· | ···················· | ···················· |
| Y | 8266 | hCoV-19/England/205090260/2020\|EPI_ISL_728343\|2020 | ···················· | ···················· | ···················· | ···················· | ···················· | ···················· |
| Y | 8267 | hCoV-19/Morocco/FMP-22/2020\|EPI_ISL_728344\|2020-12 | ···················· | ···················· | ···················· | ···················· | ···················· | ···················· |
| Y | 8268 | hCoV-19/Morocco/FMP-23/2020\|EPI_ISL_728347\|2020-12 | ···················· | ···················· | ···················· | ···················· | ···················· | ···················· |
| Y | 8269 | hCoV-19/Morocco/FMP-25/2020\|EPI_ISL_728352\|2020-12 | ···················· | ···················· | ···················· | ···················· | ···················· | ···················· |
| Y | 8270 | hCoV-19/Morocco/FMP-26/2020\|EPI_ISL_728353\|2020-12 | ···················· | ···················· | ···················· | ···················· | ···················· | ···················· |
| Y | 8271 | hCoV-19/Morocco/FMP-27/2020\|EPI_ISL_728355\|2020-12 | ···················· | ···················· | ···················· | ···················· | ···················· | ···················· |
| Y | 8272 | hCoV-19/USA/MI-UM-10036782516/2020\|EPI_ISL_728370\| | ···················· | ···················· | ···················· | ···················· | ···················· | ···················· |
| Y | 8273 | hCoV-19/USA/MI-UM-10036779751/2020\|EPI_ISL_728371\| | ···················· | ···················· | ···················· | ···················· | ···················· | ···················· |
| Y | 8274 | hCoV-19/USA/MI-UM-10036779118/2020\|EPI_ISL_728373\| | ···················· | ···················· | ···················· | ···················· | ···················· | ···················· |
| Y | 8277 | hCoV-19/USA/MI-UM-10036817031/2020\|EPI_ISL_728376\| | ···················· | ···················· | ···················· | ···················· | ···················· | ···················· |
| Y | 8278 | hCoV-19/USA/MI-UM-10036815482/2020\|EPI_ISL_728377\| | ···················· | ···················· | ···················· | ···················· | ···················· | ···················· |
| Y | 8279 | hCoV-19/USA/MI-UM-10036821132/2020\|EPI_ISL_728378\| | ···················· | ···················· | ···················· | ···················· | ···················· | ···················· |
| Y | 8280 | hCoV-19/USA/MI-UM-10036825721/2020\|EPI_ISL_728379\| | ···················· | ···················· | ···················· | ···················· | ···················· | ···················· |
| Y | 8281 | hCoV-19/USA/MI-UM-10036799382/2020\|EPI_ISL_728380\| | ···················· | ···················· | ···················· | ···················· | ···················· | ···················· |
| Y | 8282 | hCoV-19/USA/MI-UM-10036802807/2020\|EPI_ISL_728381\| | ···················· | ···················· | ···················· | ···················· | ···················· | ···················· |
| Y | 8283 | hCoV-19/USA/MI-UM-10036814492/2020\|EPI_ISL_728382\| | ···················· | ···················· | ···················· | ···················· | ···················· | ···················· |
| Y | 8284 | hCoV-19/USA/MI-UM-10036826833/2020\|EPI_ISL_728383\| | ···················· | ···················· | ···················· | ···················· | ···················· | ···················· |
| Y | 8285 | hCoV-19/USA/MI-UM-10036806081/2020\|EPI_ISL_728384\| | ···················· | ···················· | ···················· | ···················· | ···················· | ···················· |
| Y | 8286 | hCoV-19/USA/MI-UM-10036817592/2020\|EPI_ISL_728385\| | ···················· | ···················· | ···················· | ···················· | ···················· | ···················· |
| Y | 8287 | hCoV-19/USA/MI-UM-10036823317/2020\|EPI_ISL_728386\| | ···················· | ···················· | ···················· | ···················· | ···················· | ···················· |
| Y | 8288 | hCoV-19/USA/MI-UM-10036813767/2020\|EPI_ISL_728387\| | ···················· | ···················· | ···················· | ···················· | ···················· | ···················· |
| Y | 8289 | hCoV-19/USA/MI-UM-10036806101/2020\|EPI_ISL_728388\| | ···················· | ······A············· | ···················· | ···················· | ···················· | ···················· |
| Y | 8290 | hCoV-19/USA/MI-UM-10036817617/2020\|EPI_ISL_728389\| | ···················· | ···················· | ···················· | ···················· | ···················· | ···················· |
| Y | 8292 | hCoV-19/Germany/DE-BE-ChVir9004/2020\|EPI_ISL_72960 | ···················· | ···················· | ···················· | ···················· | ···················· | ···················· |
| Y | 8293 | hCoV-19/Germany/DE-BE-ChVir9006/2020\|EPI_ISL_72960 | ···················· | ···················· | ···················· | ···················· | ···················· | ···················· |
| Y | 8294 | hCoV-19/Germany/DE-BE-ChVir9009/2020\|EPI_ISL_72960 | ···················· | ···················· | ···················· | ···················· | ···················· | ···················· |
| Y | 8295 | hCoV-19/Germany/DE-BE-ChVir9012/2020\|EPI_ISL_72960 | ···················· | ·A····A············· | ···················· | ···················· | ···················· | ···················· |
| Y | 8296 | hCoV-19/Germany/DE-BE-ChVir9029/2020\|EPI_ISL_72960 | ···················· | ·A····A············· | ···················· | ···················· | ···················· | ···················· |
| Y | 8297 | hCoV-19/Germany/DE-BE-ChVir9030/2020\|EPI_ISL_72960 | ···················· | ···················· | ···················· | ···················· | ···················· | ···················· |
| Y | 8298 | hCoV-19/Germany/DE-BE-ChVir9034/2020\|EPI_ISL_72960 | ···················· | ···················· | ···················· | ···················· | ···················· | ···················· |
| Y | 8299 | hCoV-19/Germany/DE-SH-ChVir9120/2020\|EPI_ISL_72960 | ···················· | ···················· | ···················· | ···················· | ···················· | ···················· |
| Y | 8300 | hCoV-19/Germany/DE-SH-ChVir9124/2020\|EPI_ISL_72960 | ···················· | ···················· | ···················· | ···················· | ···················· | ···················· |
| Y | 8301 | hCoV-19/Germany/DE-SH-ChVir20302/2020\|EPI_ISL_7296 | ···················· | ···················· | ···················· | ···················· | ···················· | ···················· |
| Y | 8302 | hCoV-19/Germany/ChVir20304/2020\|EPI_ISL_729641\|202 | ···················· | ···················· | ···················· | ···················· | ···················· | ···················· |
| Y | 8303 | hCoV-19/Germany/DE-SH-ChVir20305/2020\|EPI_ISL_7296 | ···················· | ···················· | ···················· | ···················· | ···················· | ···················· |
| Y | 8304 | hCoV-19/Germany/DE-SH-ChVir20306/2020\|EPI_ISL_7296 | ···················· | ···················· | ···················· | ···················· | ···················· | ···················· |
| Y | 8305 | hCoV-19/Germany/DE-SH-ChVir20307/2020\|EPI_ISL_7296 | ···················· | ···················· | ···················· | ···················· | ···················· | ···················· |
| Y | 8306 | hCoV-19/Germany/DE-SH-ChVir20308/2020\|EPI_ISL_7296 | ···················· | ···················· | ···················· | ···················· | ···················· | ···················· |

| | | | | | | | | |
|---|---|---|---|---|---|---|---|---|
| Y | 8307 | hCoV-19/Germany/ChVir20311/2020\|EPI_ISL_729646\|202 | ···················· | ···················· | ···················· | ···················· | ···················· | ···················· |
| Y | 8308 | hCoV-19/Germany/ChVir20312/2020\|EPI_ISL_729647\|202 | ···················· | ···················· | ···················· | ···················· | ···················· | ···················· |
| Y | 8309 | hCoV-19/Germany/DE-SH-ChVir20313/2020\|EPI_ISL_7296 | ···················· | ···················· | ···················· | ···················· | ···················· | ···················· |
| Y | 8310 | hCoV-19/Germany/ChVir20314/2020\|EPI_ISL_729649\|202 | ···················· | ···················· | ···················· | ···················· | ···················· | ···················· |
| Y | 8311 | hCoV-19/Germany/DE-SH-ChVir20315/2020\|EPI_ISL_7296 | ···················· | ···················· | ···················· | ···················· | ···················· | ···················· |
| Y | 8312 | hCoV-19/Germany/DE-HH-ChVir20316/2020\|EPI_ISL_7296 | ···················· | ···················· | ···················· | ···················· | ···················· | ···················· |
| Y | 8313 | hCoV-19/Germany/ChVir20318/2020\|EPI_ISL_729652\|202 | ···················· | ···················· | ···················· | ···················· | ···················· | ···················· |
| Y | 8314 | hCoV-19/Germany/DE-SH-ChVir20291/2020\|EPI_ISL_7296 | ···················· | ···················· | ···················· | ···················· | ···················· | ···················· |
| Y | 8315 | hCoV-19/Germany/DE-SH-ChVir20292/2020\|EPI_ISL_7296 | ···················· | ···················· | ···················· | ···················· | ···················· | ···················· |
| Y | 8316 | hCoV-19/Germany/DE-SH-ChVir20294/2020\|EPI_ISL_7296 | ···················· | ···················· | ···················· | ···················· | ···················· | ···················· |
| Y | 8317 | hCoV-19/Germany/DE-SH-ChVir20295/2020\|EPI_ISL_7296 | ···················· | ···················· | ···················· | ···················· | ···················· | ···················· |
| Y | 8318 | hCoV-19/Germany/DE-SH-ChVir9978/2020\|EPI_ISL_72965 | ···················· | ·A·················· | ···················· | ···················· | ···················· | ···················· |
| Y | 8319 | hCoV-19/Germany/DE-SH-ChVir9994/2020\|EPI_ISL_72965 | ···················· | ···················· | ···················· | ···················· | ···················· | ···················· |
| Y | 8320 | hCoV-19/Germany/DE-SH-ChVir20028/2020\|EPI_ISL_7296 | ···················· | ···················· | ···················· | ···················· | ···················· | ···················· |
| Y | 8323 | hCoV-19/Germany/DE-SH-ChVir20047/2020\|EPI_ISL_7296 | ···················· | ···A················ | ···················· | ···················· | ···················· | ···················· |
| Y | 8324 | hCoV-19/Germany/DE-SH-ChVir20048/2020\|EPI_ISL_7296 | ···················· | ···A················ | ···················· | ···················· | ···················· | ···················· |
| Y | 8325 | hCoV-19/Germany/DE-SH-ChVir9977/2020\|EPI_ISL_72966 | ···················· | ···················· | ···················· | ···················· | ······T············· | ···················· |
| Y | 8326 | hCoV-19/Germany/DE-SH-ChVir9991/2020\|EPI_ISL_72966 | ···················· | ···················· | ···················· | ···················· | ···················· | ···················· |
| Y | 8327 | hCoV-19/Germany/DE-SH-ChVir9993/2020\|EPI_ISL_72966 | ···················· | ···················· | ···················· | ···················· | ······T············· | ···················· |
| Y | 8328 | hCoV-19/Germany/DE-SH-ChVir9999/2020\|EPI_ISL_72966 | ···················· | ···················· | ···················· | ···················· | ···················· | ···················· |
| Y | 8329 | hCoV-19/Germany/DE-SH-ChVir20056/2020\|EPI_ISL_7296 | ···················· | ···················· | ···················· | ···················· | ···················· | ···················· |
| Y | 8330 | hCoV-19/Germany/DE-SH-ChVir20000/2020\|EPI_ISL_7296 | ···················· | ···················· | ···················· | ···················· | ···················· | ···················· |
| Y | 8331 | hCoV-19/Germany/DE-SH-ChVir20050/2020\|EPI_ISL_7296 | ···················· | ···················· | ···················· | ···················· | ···················· | ···················· |
| Y | 8332 | hCoV-19/Germany/DE-SH-ChVir20728/2020\|EPI_ISL_7296 | ···················· | ···················· | ···················· | ···················· | ···················· | ···················· |
| Y | 8333 | hCoV-19/Germany/DE-SH-ChVir20729/2020\|EPI_ISL_7296 | ···················· | ···················· | ···················· | ···················· | ···················· | ···················· |
| Y | 8334 | hCoV-19/Germany/DE-BY-ChVir20731/2020\|EPI_ISL_7296 | ···················· | ···················· | ···················· | ···················· | ···················· | ···················· |
| Y | 8335 | hCoV-19/Germany/DE-SH-ChVir20734/2020\|EPI_ISL_7296 | ···················· | ···················· | ···················· | ···················· | ···················· | ···················· |
| Y | 8336 | hCoV-19/Germany/DE-SH-ChVir20735/2020\|EPI_ISL_7296 | ···················· | ···················· | ···················· | ···················· | ···················· | ···················· |
| Y | 8337 | hCoV-19/Germany/DE-SH-ChVir20736/2020\|EPI_ISL_7296 | ···················· | ···················· | ···················· | ···················· | ···················· | ···················· |
| Y | 8338 | hCoV-19/Germany/DE-SH-ChVir20737/2020\|EPI_ISL_7296 | ···················· | ···················· | ···················· | ···················· | ···················· | ···················· |
| Y | 8339 | hCoV-19/Germany/DE-SH-ChVir20739/2020\|EPI_ISL_7296 | ···················· | ···················· | ···················· | ···················· | ···················· | ···················· |
| Y | 8340 | hCoV-19/Germany/DE-SH-ChVir20740/2020\|EPI_ISL_7296 | ···················· | ···················· | ···················· | ···················· | ···················· | ···················· |
| Y | 8341 | hCoV-19/Germany/DE-SH-ChVir20780/2020\|EPI_ISL_7296 | ···················· | ···················· | ···················· | ···················· | ···················· | ···················· |
| Y | 8342 | hCoV-19/Germany/DE-SH-ChVir20781/2020\|EPI_ISL_7296 | ···················· | ·A·················· | ···················· | ···················· | ···················· | ···················· |
| Y | 8343 | hCoV-19/Germany/DE-SH-ChVir20782/2020\|EPI_ISL_7296 | ···················· | ···················· | ···················· | ···················· | ···················· | ···················· |
| Y | 8344 | hCoV-19/Germany/DE-SH-ChVir20783/2020\|EPI_ISL_7296 | ···················· | ···················· | ···················· | ···················· | ···················· | ···················· |
| Y | 8345 | hCoV-19/Switzerland/SG-UZH-IMV124/2020\|EPI_ISL_730 | ···················· | ···················· | ···················· | ···················· | ···················· | ···················· |
| Y | 8346 | hCoV-19/Switzerland/TG-UZH-IMV116/2020\|EPI_ISL_730 | ···················· | ···················· | ···················· | ···················· | ···················· | ···················· |
| Y | 8347 | hCoV-19/Switzerland/ZH-UZH-IMV107/2020\|EPI_ISL_730 | ···················· | ···················· | ···················· | ···················· | ···················· | ···················· |
| Y | 8348 | hCoV-19/Switzerland/ZH-UZH-IMV108/2020\|EPI_ISL_730 | ···················· | ···················· | ···················· | ···················· | ···················· | ···················· |
| Y | 8349 | hCoV-19/Switzerland/ZH-UZH-IMV109/2020\|EPI_ISL_730 | ···················· | ···················· | ···················· | ···················· | ···················· | ···················· |
| Y | 8350 | hCoV-19/Switzerland/ZH-UZH-IMV110/2020\|EPI_ISL_730 | ···················· | ···················· | ···················· | ···················· | ···················· | ···················· |
| Y | 8351 | hCoV-19/Switzerland/ZH-UZH-IMV111/2020\|EPI_ISL_730 | ···················· | ···················· | ···················· | ···················· | ···················· | ···················· |
| Y | 8352 | hCoV-19/Switzerland/ZH-UZH-IMV112/2020\|EPI_ISL_730 | ···················· | ···················· | ···················· | ···················· | ···················· | ···················· |
| Y | 8353 | hCoV-19/Switzerland/ZH-UZH-IMV113/2020\|EPI_ISL_730 | ···················· | ···················· | ···················· | ···················· | ···················· | ···················· |
| Y | 8354 | hCoV-19/Switzerland/ZH-UZH-IMV114/2020\|EPI_ISL_730 | ···················· | ···················· | ···················· | ···················· | ···················· | ···················· |
| Y | 8355 | hCoV-19/Switzerland/ZH-UZH-IMV115/2020\|EPI_ISL_730 | ···················· | ···················· | ···········Y········ | ···················· | ···················· | ···················· |
| Y | 8356 | hCoV-19/Switzerland/ZH-UZH-IMV117/2020\|EPI_ISL_730 | ···················· | ···················· | ···················· | ···················· | ···················· | ···················· |
| Y | 8357 | hCoV-19/Switzerland/ZH-UZH-IMV118/2020\|EPI_ISL_730 | ···················· | ···················· | ···················· | ···················· | ···················· | ···················· |
| Y | 8358 | hCoV-19/Switzerland/ZH-UZH-IMV119/2020\|EPI_ISL_730 | ···················· | ···················· | ···················· | ···················· | ···················· | ···················· |
| Y | 8359 | hCoV-19/Switzerland/ZH-UZH-IMV120/2020\|EPI_ISL_730 | ···················· | ···················· | ···················· | ···················· | ···················· | ···················· |
| Y | 8360 | hCoV-19/Switzerland/ZH-UZH-IMV121/2020\|EPI_ISL_730 | ···················· | ···················· | ···················· | ···················· | ···················· | ···················· |
| Y | 8361 | hCoV-19/Switzerland/ZH-UZH-IMV123/2020\|EPI_ISL_730 | ···················· | ···················· | ···················· | ···················· | ···················· | ···················· |
| Y | 8362 | hCoV-19/Switzerland/ZH-UZH-IMV125/2020\|EPI_ISL_730 | ···················· | ···················· | ···················· | ···················· | ···················· | ···················· |
| Y | 8363 | hCoV-19/France/ARA-104480/2020\|EPI_ISL_730640\|2020 | ···················· | ···················· | ···················· | ···················· | ···················· | ···················· |
| Y | 8364 | hCoV-19/France/ARA-104482/2020\|EPI_ISL_730641\|2020 | ···················· | ···················· | ···················· | ···················· | ···················· | ···················· |
| Y | 8365 | hCoV-19/France/ARA-104489/2020\|EPI_ISL_730642\|2020 | ···················· | ···················· | ···················· | ···················· | ···················· | ···················· |
| Y | 8366 | hCoV-19/France/ARA-104519/2020\|EPI_ISL_730643\|2020 | ···················· | ···················· | ···················· | ···················· | ···················· | ···················· |
| Y | 8367 | hCoV-19/France/ARA-104739/2020\|EPI_ISL_730644\|2020 | ···················· | ···················· | ···················· | ···················· | ···················· | ···················· |
| Y | 8368 | hCoV-19/France/ARA-104821/2020\|EPI_ISL_730645\|2020 | ···················· | ···················· | ···················· | ···················· | ···················· | ···················· |
| Y | 8369 | hCoV-19/France/ARA-104822/2020\|EPI_ISL_730646\|2020 | ···················· | ···················· | ···················· | ···················· | ···················· | ···················· |
| Y | 8370 | hCoV-19/France/ARA-104824/2020\|EPI_ISL_730647\|2020 | ···················· | ···················· | ···················· | ···················· | ···················· | ···················· |
| Y | 8371 | hCoV-19/France/ARA-104838/2020\|EPI_ISL_730648\|2020 | ···················· | ···················· | ···················· | ···················· | ···················· | ···················· |
| Y | 8372 | hCoV-19/France/ARA-104848/2020\|EPI_ISL_730649\|2020 | ···················· | ···················· | ···················· | ···················· | ···················· | ···················· |
| Y | 8373 | hCoV-19/France/ARA-105054/2020\|EPI_ISL_730650\|2020 | ···················· | ···················· | ···················· | ···················· | ···················· | ···················· |
| Y | 8374 | hCoV-19/France/ARA-105237/2020\|EPI_ISL_730651\|2020 | ···················· | ···················· | ···················· | ···················· | ···················· | ···················· |
| Y | 8375 | hCoV-19/dog/Italy/Dog399-20BA/2020\|EPI_ISL_730652\| | ···················· | ···················· | ···················· | ···················· | ···················· | ···················· |
| Y | 8376 | hCoV-19/Italy/TE.237004/2020\|EPI_ISL_730653\|2020-1 | ···················· | ···················· | ···················· | ···················· | ···················· | ···················· |
| Y | 8377 | hCoV-19/England/QEUH-C3227C/2020\|EPI_ISL_730656\|20 | ···················· | ···················· | ···················· | ···················· | ···················· | ···················· |
| Y | 8378 | hCoV-19/England/QEUH-C319B4/2020\|EPI_ISL_730657\|20 | ···················· | ···················· | ···················· | ···················· | ···················· | ···················· |
| Y | 8379 | hCoV-19/England/ALDP-C3D903/2020\|EPI_ISL_730658\|20 | ···················· | ···················· | ···················· | ···················· | ···················· | ···················· |
| Y | 8380 | hCoV-19/England/QEUH-C32203/2020\|EPI_ISL_730659\|20 | ···················· | ···················· | ···················· | ···················· | ···················· | ···················· |
| Y | 8381 | hCoV-19/England/ALDP-C3AE34/2020\|EPI_ISL_730660\|20 | ···················· | ···················· | ···················· | ···················· | ···················· | ···················· |
| Y | 8382 | hCoV-19/England/ALDP-C3AEE9/2020\|EPI_ISL_730663\|20 | ···················· | ·A·················· | ···················· | ···················· | ···················· | ···A················ |
| Y | 8383 | hCoV-19/England/QEUH-C30F68/2020\|EPI_ISL_730664\|20 | ···················· | ···················· | ···················· | ···················· | ···················· | ···················· |
| Y | 8384 | hCoV-19/England/ALDP-C3B659/2020\|EPI_ISL_730665\|20 | ···················· | ···················· | ···················· | ···················· | ···················· | ···················· |
| Y | 8385 | hCoV-19/England/QEUH-C31A1B/2020\|EPI_ISL_730678\|20 | ···················· | ···················· | ···················· | ···················· | ···················· | ···················· |
| Y | 8386 | hCoV-19/England/ALDP-C39D38/2020\|EPI_ISL_730679\|20 | ···················· | ···················· | ···················· | ···················· | ···················· | ···················· |
| Y | 8387 | hCoV-19/England/ALDP-C3DBD0/2020\|EPI_ISL_730682\|20 | ···················· | ···················· | ···················· | ···················· | ···················· | ···················· |
| Y | 8388 | hCoV-19/England/ALDP-C3B1C1/2020\|EPI_ISL_730688\|20 | ···················· | ···················· | ···················· | ···················· | ···················· | ···················· |

| | | | | | | | | |
|---|---|---|---|---|---|---|---|---|
| Y | 8389 | hCoV-19/England/ALDP-C39688/2020\|EPI_ISL_730689\|20 | ···················· | ···················· | ···················· | ···················· | ···················· | ···················· |
| Y | 8390 | hCoV-19/England/ALDP-C39CFF/2020\|EPI_ISL_730690\|20 | ···················· | ···················· | ···················· | ···················· | ···················· | ···················· |
| Y | 8391 | hCoV-19/England/ALDP-C3DDAD/2020\|EPI_ISL_730692\|20 | ···················· | ···················· | ···················· | ···················· | ···················· | ···················· |
| Y | 8392 | hCoV-19/England/ALDP-C3B404/2020\|EPI_ISL_730693\|20 | ···················· | ···················· | ···················· | ···················· | ···················· | ···················· |
| Y | 8393 | hCoV-19/England/ALDP-C3DBA3/2020\|EPI_ISL_730696\|20 | ···················· | ···················· | ···················· | ···················· | ···················· | ···················· |
| Y | 8394 | hCoV-19/England/QEUH-C30DD7/2020\|EPI_ISL_730697\|20 | ···················· | ···················· | ···················· | ···················· | ···················· | ···················· |
| Y | 8395 | hCoV-19/England/ALDP-C39B01/2020\|EPI_ISL_730698\|20 | ···················· | ···················· | ···················· | ···················· | ······T············· | ···················· |
| Y | 8396 | hCoV-19/England/ALDP-C3D851/2020\|EPI_ISL_730699\|20 | ···················· | ···················· | ···················· | ···················· | ···················· | ···················· |
| Y | 8397 | hCoV-19/Portugal/PT1648/2020\|EPI_ISL_731953\|2020-1 | ···················· | ···················· | ···················· | ···················· | ···················· | ···················· |
| Y | 8398 | hCoV-19/Portugal/PT1649/2020\|EPI_ISL_731954\|2020-1 | ···················· | ···················· | ···················· | ···················· | ···················· | ···················· |
| Y | 8399 | hCoV-19/Portugal/PT1651/2020\|EPI_ISL_731955\|2020-1 | ···················· | ···················· | ···················· | ···················· | ···················· | ···················· |
| Y | 8400 | hCoV-19/Portugal/PT1654/2020\|EPI_ISL_731956\|2020-1 | ···················· | ···················· | ···················· | ···················· | ···················· | ···················· |
| Y | 8401 | hCoV-19/Portugal/PT1655/2020\|EPI_ISL_731957\|2020-1 | ···················· | ···················· | ···················· | ···················· | ···················· | ···················· |
| Y | 8402 | hCoV-19/Portugal/PT1656/2020\|EPI_ISL_731958\|2020-1 | ···················· | ···················· | ···················· | ···················· | ···················· | ···················· |
| Y | 8403 | hCoV-19/Portugal/PT1657/2020\|EPI_ISL_731959\|2020-1 | ···················· | ···················· | ···················· | ···················· | ···················· | ···················· |
| Y | 8404 | hCoV-19/Portugal/PT1658/2020\|EPI_ISL_731960\|2020-1 | ···················· | ···················· | ···················· | ···················· | ···················· | ···················· |
| Y | 8405 | hCoV-19/Portugal/PT1659/2020\|EPI_ISL_731961\|2020-1 | ···················· | ···················· | ···················· | ···················· | ···················· | ···················· |
| Y | 8406 | hCoV-19/Portugal/PT1660/2020\|EPI_ISL_731962\|2020-1 | ···················· | ···················· | ···················· | ···················· | ···················· | ···················· |
| Y | 8407 | hCoV-19/Portugal/PT1661/2020\|EPI_ISL_731963\|2020-1 | ···················· | ···················· | ···················· | ···················· | ···················· | ···················· |
| Y | 8408 | hCoV-19/Portugal/PT1663/2020\|EPI_ISL_731964\|2020-1 | ···················· | ···················· | ···················· | ···················· | ···················· | ···················· |
| Y | 8409 | hCoV-19/Portugal/PT1664/2020\|EPI_ISL_731965\|2020-1 | ···················· | ···················· | ···················· | ···················· | ···················· | ···················· |
| Y | 8410 | hCoV-19/Portugal/PT1667/2020\|EPI_ISL_731966\|2020-1 | ···················· | ···················· | ···················· | ···················· | ···················· | ···················· |
| Y | 8411 | hCoV-19/Portugal/PT1668/2020\|EPI_ISL_731967\|2020-1 | ···················· | ···················· | ···················· | ···················· | ···················· | ···················· |
| Y | 8412 | hCoV-19/Portugal/PT1669/2020\|EPI_ISL_731968\|2020-1 | ···················· | ···················· | ···················· | ···················· | ···················· | ···················· |
| Y | 8413 | hCoV-19/Portugal/PT1670/2020\|EPI_ISL_731969\|2020-1 | ···················· | ···················· | ···················· | ···················· | ···················· | ···················· |
| Y | 8414 | hCoV-19/Portugal/PT1672/2020\|EPI_ISL_731970\|2020-1 | ···················· | ···················· | ···················· | ···················· | ···················· | ···················· |
| Y | 8415 | hCoV-19/Portugal/PT1673/2020\|EPI_ISL_731971\|2020-1 | ···················· | ···················· | ···················· | ···················· | ···················· | ···················· |
| Y | 8416 | hCoV-19/Portugal/PT1674/2020\|EPI_ISL_731972\|2020-1 | ···················· | ···················· | ···················· | ···················· | ···················· | ···················· |
| Y | 8417 | hCoV-19/Portugal/PT1675/2020\|EPI_ISL_731973\|2020-1 | ···················· | ···················· | ···················· | ···················· | ···················· | ···················· |
| Y | 8418 | hCoV-19/Portugal/PT1676/2020\|EPI_ISL_731974\|2020-1 | ···················· | ···················· | ···················· | ···················· | ···················· | ···················· |
| Y | 8419 | hCoV-19/Portugal/PT1678/2020\|EPI_ISL_731975\|2020-1 | ···················· | ···················· | ···················· | ···················· | ···················· | ···················· |
| Y | 8420 | hCoV-19/Portugal/PT1679/2020\|EPI_ISL_731976\|2020-1 | ···················· | ···················· | ···················· | ···················· | ···················· | ···················· |
| Y | 8421 | hCoV-19/Portugal/PT1740/2020\|EPI_ISL_731977\|2020-1 | ···················· | ···················· | ···················· | ···················· | ···················· | ···················· |
| Y | 8422 | hCoV-19/Portugal/PT1741/2020\|EPI_ISL_731978\|2020-1 | ···················· | ···················· | ···················· | ···················· | ···················· | ···················· |
| Y | 8423 | hCoV-19/Portugal/PT1742/2020\|EPI_ISL_731979\|2020-1 | ···················· | ···················· | ···················· | ···················· | ···················· | ···················· |
| Y | 8424 | hCoV-19/Portugal/PT1743/2020\|EPI_ISL_731980\|2020-1 | ···················· | ···················· | ···················· | ···················· | ···················· | ···················· |
| Y | 8425 | hCoV-19/Portugal/PT1744/2020\|EPI_ISL_731981\|2020-1 | ···················· | ···················· | ···················· | ···················· | ···················· | ···················· |
| Y | 8426 | hCoV-19/Portugal/PT1745/2020\|EPI_ISL_731982\|2020-1 | ···················· | ···················· | ···················· | ···················· | ···················· | ···················· |
| Y | 8427 | hCoV-19/Portugal/PT1746/2020\|EPI_ISL_731983\|2020-1 | ···················· | ···················· | ···················· | ···················· | ···················· | ···················· |
| Y | 8428 | hCoV-19/Portugal/PT1747/2020\|EPI_ISL_731984\|2020-1 | ···················· | ···················· | ···················· | ···················· | ···················· | ···················· |
| Y | 8429 | hCoV-19/Portugal/PT1748/2020\|EPI_ISL_731985\|2020-1 | ···················· | ···················· | ···················· | ···················· | ···················· | ···················· |
| Y | 8430 | hCoV-19/Portugal/PT1749/2020\|EPI_ISL_731986\|2020-1 | ···················· | ···················· | ···················· | ···················· | ···················· | ···················· |
| Y | 8431 | hCoV-19/Portugal/PT1750/2020\|EPI_ISL_731987\|2020-1 | ···················· | ···················· | ···················· | ···················· | ···················· | ···················· |
| Y | 8432 | hCoV-19/Portugal/PT1751/2020\|EPI_ISL_731988\|2020-1 | ···················· | ···················· | ···················· | ···················· | ···················· | ···················· |
| Y | 8433 | hCoV-19/Portugal/PT1753/2020\|EPI_ISL_731989\|2020-1 | ···················· | ···················· | ···················· | ···················· | ···················· | ···················· |
| Y | 8434 | hCoV-19/Portugal/PT1754/2020\|EPI_ISL_731990\|2020-1 | ···················· | ···················· | ···················· | ···················· | ···················· | ···················· |
| Y | 8435 | hCoV-19/Portugal/PT1755/2020\|EPI_ISL_731991\|2020-1 | ···················· | ···················· | ···················· | ···················· | ···················· | ···················· |
| Y | 8436 | hCoV-19/Portugal/PT1756/2020\|EPI_ISL_731992\|2020-1 | ···················· | ···················· | ···················· | ···················· | ···················· | ···················· |
| Y | 8437 | hCoV-19/Portugal/PT1759/2020\|EPI_ISL_731993\|2020-1 | ···················· | ···················· | ···················· | ···················· | ···················· | ···················· |
| Y | 8438 | hCoV-19/Portugal/PT1760/2020\|EPI_ISL_731994\|2020-1 | ···················· | ···················· | ···················· | ···················· | ···················· | ···················· |
| Y | 8439 | hCoV-19/Portugal/PT1761/2020\|EPI_ISL_731995\|2020-1 | ···················· | ···················· | ···················· | ···················· | ···················· | ···················· |
| Y | 8440 | hCoV-19/Portugal/PT1762/2020\|EPI_ISL_731996\|2020-1 | ···················· | ···················· | ···················· | ···················· | ···················· | ···················· |
| Y | 8441 | hCoV-19/USA/VA-DCLS-2443/2020\|EPI_ISL_734192\|2020- | ···················· | ······A············· | ···················· | ···················· | ···················· | ···················· |
| Y | 8442 | hCoV-19/USA/VA-DCLS-2444/2020\|EPI_ISL_734193\|2020- | ···················· | ···················· | ···················· | ···················· | ···················· | ···················· |
| Y | 8443 | hCoV-19/USA/VA-DCLS-2447/2020\|EPI_ISL_734194\|2020- | ···················· | ···················· | ···················· | ···················· | ···················· | ···················· |
| Y | 8444 | hCoV-19/USA/VA-DCLS-2448/2020\|EPI_ISL_734195\|2020- | ···················· | ···················· | ···················· | ···················· | ···················· | ···················· |
| Y | 8445 | hCoV-19/USA/VA-DCLS-2449/2020\|EPI_ISL_734196\|2020- | ···················· | ···················· | ···················· | ···················· | ···················· | ···················· |
| Y | 8446 | hCoV-19/USA/VA-DCLS-2450/2020\|EPI_ISL_734197\|2020- | ···················· | ···················· | ···················· | ···················· | ···················· | ···················· |
| Y | 8447 | hCoV-19/USA/VA-DCLS-2453/2020\|EPI_ISL_734198\|2020- | ···················· | ···················· | ···················· | ···················· | ···················· | ···················· |
| Y | 8448 | hCoV-19/USA/VA-DCLS-2454/2020\|EPI_ISL_734199\|2020- | ···················· | ···················· | ···················· | ···················· | ···················· | ···················· |
| Y | 8449 | hCoV-19/England/MILK-B877D4/2020\|EPI_ISL_724800\|20 | ···················· | ···················· | ···················· | ···················· | ······T············· | ···················· |
| Y | 8450 | hCoV-19/England/MILK-B877E3/2020\|EPI_ISL_724801\|20 | ···················· | ···················· | ···················· | ···················· | ···················· | ···················· |
| Y | 8451 | hCoV-19/England/MILK-B877F2/2020\|EPI_ISL_724802\|20 | ···················· | ···················· | ···················· | ···················· | ···················· | ···················· |
| Y | 8452 | hCoV-19/England/MILK-B8780E/2020\|EPI_ISL_724803\|20 | ···················· | ···················· | ···················· | ···················· | ···················· | ···················· |
| Y | 8453 | hCoV-19/England/MILK-B8783B/2020\|EPI_ISL_724805\|20 | ···········T········ | ···················· | ···················· | ···················· | ···················· | ···················· |
| Y | 8454 | hCoV-19/England/MILK-B87886/2020\|EPI_ISL_724808\|20 | ···················· | ···················· | ···················· | ···················· | ···················· | ···················· |
| Y | 8455 | hCoV-19/England/MILK-B87895/2020\|EPI_ISL_724809\|20 | ···················· | ···················· | ···················· | ···················· | ···················· | ···················· |
| Y | 8456 | hCoV-19/England/MILK-B878A4/2020\|EPI_ISL_724810\|20 | ···················· | ···················· | ···················· | ···················· | ···················· | ···················· |
| Y | 8457 | hCoV-19/England/MILK-B878C2/2020\|EPI_ISL_724811\|20 | ···················· | ···················· | ···················· | ···················· | ······T············· | ···················· |
| Y | 8458 | hCoV-19/England/MILK-B878FF/2020\|EPI_ISL_724812\|20 | ···················· | ···················· | ···················· | ···················· | ···················· | ···················· |
| Y | 8459 | hCoV-19/England/MILK-B87929/2020\|EPI_ISL_724813\|20 | ···················· | ···················· | ···················· | ···················· | ···················· | ···················· |
| Y | 8460 | hCoV-19/England/MILK-B87938/2020\|EPI_ISL_724814\|20 | ···················· | ···················· | ···················· | ···················· | ···················· | ···················· |
| Y | 8461 | hCoV-19/England/MILK-B87992/2020\|EPI_ISL_724816\|20 | ···················· | ···················· | ···················· | ···················· | ······T············· | ···················· |
| Y | 8462 | hCoV-19/England/MILK-B879FC/2020\|EPI_ISL_724820\|20 | ···················· | ···················· | ···················· | ···················· | ···················· | ···················· |
| Y | 8463 | hCoV-19/England/MILK-B87A17/2020\|EPI_ISL_724821\|20 | ···················· | ···················· | ···················· | ···················· | ···················· | ···················· |
| Y | 8464 | hCoV-19/England/MILK-B87A26/2020\|EPI_ISL_724822\|20 | ···················· | ···················· | ···················· | ···················· | ···················· | ···················· |
| Y | 8465 | hCoV-19/England/MILK-B87A44/2020\|EPI_ISL_724824\|20 | ···················· | ···················· | ···················· | ···················· | ······T············· | ···················· |
| Y | 8466 | hCoV-19/England/MILK-B87A71/2020\|EPI_ISL_724825\|20 | ···················· | ···················· | ···················· | ···················· | ······T············· | ···················· |
| Y | 8467 | hCoV-19/England/MILK-B87A9F/2020\|EPI_ISL_724826\|20 | ···················· | ···················· | ···················· | ···················· | ···················· | ···················· |
| Y | 8468 | hCoV-19/England/MILK-B87AEA/2020\|EPI_ISL_724830\|20 | ···················· | ·A·················· | ···················· | ···················· | ···················· | ···················· |

| | | | | | | | |
|---|---|---|---|---|---|---|---|
| Y | 8469 hCoV-19/England/MILK-B87B05/2020\|EPI_ISL_724832\|20 | ···················· | ···················· | ····················· | ···················· | ···················· | ···················· |
| Y | 8470 hCoV-19/England/MILK-B87B23/2020\|EPI_ISL_724833\|20 | ···················· | ···················· | ····················· | ···················· | ···················· | ···················· |
| Y | 8471 hCoV-19/England/MILK-B87B41/2020\|EPI_ISL_724834\|20 | ···················· | ···················· | ····················· | ···················· | ···················· | ···················· |
| Y | 8472 hCoV-19/England/MILK-B87B50/2020\|EPI_ISL_724835\|20 | ···················· | ···················· | ····················· | ···················· | ···················· | ···················· |
| Y | 8473 hCoV-19/England/NORT-1B72171/2020\|EPI_ISL_724856\|2 | ···················· | ···················· | ····················· | ···················· | ···················· | ···················· |
| Y | 8474 hCoV-19/England/NORT-1B721BD/2020\|EPI_ISL_724857\|2 | ···················· | ···················· | ····················· | ···················· | ···················· | ···················· |
| Y | 8475 hCoV-19/England/NORT-1B721EA/2020\|EPI_ISL_724859\|2 | ···················· | ···················· | ····················· | ···················· | ···················· | ···················· |
| Y | 8476 hCoV-19/England/NORT-1B72214/2020\|EPI_ISL_724860\|2 | ···················· | ···················· | ····················· | ···················· | ···················· | ···················· |
| Y | 8477 hCoV-19/England/NORT-1B7227E/2020\|EPI_ISL_724863\|2 | ···················· | ···················· | ····················· | ···················· | ···················· | ···················· |
| Y | 8478 hCoV-19/England/NORT-1B7229C/2020\|EPI_ISL_724864\|2 | ···················· | ···················· | ····················· | ···················· | ···················· | ···················· |
| Y | 8479 hCoV-19/England/NORT-1B722F6/2020\|EPI_ISL_724868\|2 | ···················· | ···················· | ····················· | ···················· | ······T············· | ···················· |
| Y | 8480 hCoV-19/England/NORT-1B7236C/2020\|EPI_ISL_724872\|2 | ···················· | ···················· | ····················· | ···················· | ···················· | ···················· |
| Y | 8481 hCoV-19/England/NORT-1B723B7/2020\|EPI_ISL_724873\|2 | ···················· | ···················· | ····················· | ···················· | ···················· | ···················· |
| Y | 8482 hCoV-19/England/NORT-1B723C6/2020\|EPI_ISL_724874\|2 | ···················· | ···················· | ····················· | ···················· | ···················· | ···················· |
| Y | 8483 hCoV-19/England/NORT-1B723F3/2020\|EPI_ISL_724876\|2 | ···················· | ···················· | ····················· | ···················· | ···················· | ···················· |
| Y | 8484 hCoV-19/England/NORT-1B725FD/2020\|EPI_ISL_724890\|2 | ···················· | ···················· | ····················· | ···················· | ···················· | ···················· |
| Y | 8485 hCoV-19/England/NORT-1B72654/2020\|EPI_ISL_724891\|2 | ···················· | ···················· | ····················· | ···················· | ···················· | ···················· |
| Y | 8486 hCoV-19/England/NORT-1B72663/2020\|EPI_ISL_724892\|2 | ···················· | ···················· | ····················· | ···················· | ···················· | ···················· |
| Y | 8487 hCoV-19/England/NORT-1B72690/2020\|EPI_ISL_724894\|2 | ···················· | ···················· | ····················· | ···················· | ···················· | ···················· |
| Y | 8488 hCoV-19/England/NORT-1B726CD/2020\|EPI_ISL_724895\|2 | ···················· | ···················· | ····················· | ···················· | ···················· | ···················· |
| Y | 8489 hCoV-19/England/NORT-1B726DC/2020\|EPI_ISL_724896\|2 | ···················· | ···················· | ····················· | ···················· | ···················· | ···················· |
| Y | 8490 hCoV-19/Wales/PHWC-498133/2020\|EPI_ISL_727030\|2020 | ···················· | ···················· | ····················· | ···················· | ···················· | ···················· |
| Y | 8491 hCoV-19/Wales/PHWC-498142/2020\|EPI_ISL_727031\|2020 | ···················· | ···················· | ····················· | ···················· | ···················· | ···················· |
| Y | 8494 hCoV-19/Wales/PHWC-49817F/2020\|EPI_ISL_727034\|2020 | ···················· | ···················· | ····················· | ···················· | ···················· | ···················· |
| Y | 8495 hCoV-19/Wales/PHWC-49818E/2020\|EPI_ISL_727035\|2020 | ···················· | ···················· | ····················· | ···················· | ···················· | ···················· |
| Y | 8496 hCoV-19/Wales/PHWC-49819D/2020\|EPI_ISL_727036\|2020 | ···················· | ···················· | ····················· | ···················· | ···················· | ···················· |
| Y | 8497 hCoV-19/Wales/PHWC-4981BB/2020\|EPI_ISL_727037\|2020 | ···················· | ···················· | ····················· | ···················· | ···················· | ···················· |
| Y | 8498 hCoV-19/Wales/PHWC-4981D9/2020\|EPI_ISL_727038\|2020 | ···················· | ···················· | ····················· | ···················· | ···················· | ···················· |
| Y | 8499 hCoV-19/Wales/PHWC-4981F7/2020\|EPI_ISL_727039\|2020 | ···················· | ···················· | ····················· | ···················· | ···················· | ···················· |
| Y | 8500 hCoV-19/Wales/PHWC-49835B/2020\|EPI_ISL_727060\|2020 | ···················· | ···················· | ····················· | ···················· | ···················· | ···················· |
| Y | 8501 hCoV-19/Wales/PHWC-49836A/2020\|EPI_ISL_727061\|2020 | ···················· | ···················· | ····················· | ···················· | ···················· | ···················· |
| Y | 8502 hCoV-19/Wales/PHWC-498379/2020\|EPI_ISL_727062\|2020 | ···················· | ···················· | ····················· | ···················· | ···················· | ···················· |
| Y | 8503 hCoV-19/Wales/PHWC-498388/2020\|EPI_ISL_727063\|2020 | ···················· | ···················· | ····················· | ···················· | ···················· | ···················· |
| Y | 8504 hCoV-19/Wales/PHWC-498397/2020\|EPI_ISL_727064\|2020 | ···················· | ···················· | ····················· | ···················· | ···················· | ···················· |
| Y | 8505 hCoV-19/Wales/PHWC-4983A6/2020\|EPI_ISL_727065\|2020 | ···················· | ···················· | ····················· | ···················· | ···················· | ···················· |
| Y | 8506 hCoV-19/Wales/PHWC-4983B5/2020\|EPI_ISL_727066\|2020 | ···················· | ···················· | ····················· | ···················· | ···················· | ···················· |
| Y | 8507 hCoV-19/Wales/PHWC-4983C4/2020\|EPI_ISL_727067\|2020 | ···················· | ···················· | ····················· | ···················· | ···················· | ···················· |
| Y | 8508 hCoV-19/Wales/PHWC-4983E2/2020\|EPI_ISL_727069\|2020 | ···················· | ···················· | ····················· | ···················· | ···················· | ···················· |
| Y | 8509 hCoV-19/Wales/PHWC-4983F1/2020\|EPI_ISL_727070\|2020 | ···················· | ···················· | ····················· | ···················· | ···················· | ···················· |
| Y | 8510 hCoV-19/Wales/PHWC-49841C/2020\|EPI_ISL_727072\|2020 | ···················· | ···················· | ····················· | ···················· | ···················· | ···················· |
| Y | 8511 hCoV-19/Wales/PHWC-49842B/2020\|EPI_ISL_727073\|2020 | ···················· | ···················· | ····················· | ···················· | ···················· | ···················· |
| Y | 8512 hCoV-19/Wales/PHWC-49843A/2020\|EPI_ISL_727074\|2020 | ···················· | ···················· | ····················· | ···················· | ···················· | ···················· |
| Y | 8513 hCoV-19/Wales/PHWC-498449/2020\|EPI_ISL_727075\|2020 | ···················· | ···················· | ····················· | ···················· | ···················· | ···················· |
| Y | 8514 hCoV-19/Wales/PHWC-498467/2020\|EPI_ISL_727077\|2020 | ···················· | ···················· | ····················· | ···················· | ···················· | ···················· |
| Y | 8515 hCoV-19/Wales/PHWC-498476/2020\|EPI_ISL_727078\|2020 | ···················· | ···················· | ····················· | ···················· | ···················· | ···················· |
| Y | 8516 hCoV-19/Wales/PHWC-498485/2020\|EPI_ISL_727079\|2020 | ···················· | ···················· | ····················· | ···················· | ···················· | ···················· |
| Y | 8517 hCoV-19/Wales/PHWC-498494/2020\|EPI_ISL_727080\|2020 | ···················· | ···················· | ····················· | ···················· | ···················· | ···················· |
| Y | 8518 hCoV-19/Wales/PHWC-4984A3/2020\|EPI_ISL_727081\|2020 | ···················· | ···················· | ····················· | ···················· | ···················· | ···················· |
| Y | 8519 hCoV-19/Wales/PHWC-4984B2/2020\|EPI_ISL_727082\|2020 | ···················· | ···················· | ····················· | ···················· | ···················· | ···················· |
| Y | 8520 hCoV-19/Wales/PHWC-4984C1/2020\|EPI_ISL_727083\|2020 | ···················· | ···················· | ····················· | ···················· | ···················· | ···················· |
| Y | 8521 hCoV-19/Wales/PHWC-4984FE/2020\|EPI_ISL_727085\|2020 | ···················· | ···················· | ····················· | ···················· | ···················· | ···················· |
| Y | 8522 hCoV-19/Wales/PHWC-49850A/2020\|EPI_ISL_727086\|2020 | ···················· | ···················· | ····················· | ···················· | ···················· | ···················· |
| Y | 8523 hCoV-19/Wales/PHWC-498519/2020\|EPI_ISL_727087\|2020 | ···················· | ···················· | ····················· | ···················· | ···················· | ···················· |
| Y | 8524 hCoV-19/Wales/PHWC-498528/2020\|EPI_ISL_727088\|2020 | ···················· | ···················· | ····················· | ···················· | ···················· | ···················· |
| Y | 8525 hCoV-19/Wales/PHWC-498537/2020\|EPI_ISL_727089\|2020 | ···················· | ···················· | ····················· | ···················· | ···················· | ···················· |
| Y | 8526 hCoV-19/Wales/PHWC-498546/2020\|EPI_ISL_727090\|2020 | ···················· | ···················· | ····················· | ···················· | ···················· | ···················· |
| Y | 8527 hCoV-19/Wales/PHWC-498555/2020\|EPI_ISL_727091\|2020 | ···················· | ···················· | ····················· | ···················· | ···················· | ···················· |
| Y | 8528 hCoV-19/Wales/PHWC-498564/2020\|EPI_ISL_727092\|2020 | ···················· | ···················· | ····················· | ···················· | ···················· | ···················· |
| Y | 8529 hCoV-19/Wales/PHWC-498573/2020\|EPI_ISL_727093\|2020 | ···················· | ···················· | ····················· | ···················· | ···················· | ···················· |
| Y | 8530 hCoV-19/Wales/PHWC-498591/2020\|EPI_ISL_727094\|2020 | ···················· | ······A············· | ····················· | ···················· | ···················· | ···················· |
| Y | 8532 hCoV-19/Wales/PHWC-4985DD/2020\|EPI_ISL_727098\|2020 | ···················· | ···················· | ····················· | ···················· | ···················· | ···················· |
| Y | 8534 hCoV-19/Morocco/FMP-29/2020\|EPI_ISL_728360\|2020-12 | T··············A···· | ···················· | ····················· | ···················· | ···················· | ···················· |
| Y | 8535 hCoV-19/Spain/CT-HUVH-GEST1-4/2020\|EPI_ISL_728361\| | ···················· | ···················· | ····················· | ···················· | ···················· | ···················· |
| Y | 8536 hCoV-19/Spain/CT-HUVH-GEST1-3/2020\|EPI_ISL_728362\| | ···················· | ···················· | ····················· | ···················· | ···················· | ···················· |
| Y | 8537 hCoV-19/Spain/CT-HUVH-GEST1-2/2020\|EPI_ISL_728364\| | ···················· | ···················· | ····················· | ···················· | ···················· | ···················· |
| Y | 8538 hCoV-19/Spain/CT-HUVH-GEST1-1/2020\|EPI_ISL_728365\| | ···················· | ···················· | ····················· | ···················· | ···················· | ···················· |
| Y | 8539 hCoV-19/Morocco/FMP-31/2020\|EPI_ISL_728366\|2020-12 | ···················· | ···················· | ····················· | ···················· | ···················· | ···················· |
| Y | 8540 hCoV-19/Morocco/FMP-32/2020\|EPI_ISL_728367\|2020-12 | ···················· | ···················· | ····················· | ···················· | ···················· | ···················· |
| Y | 8541 hCoV-19/USA/MI-UM-10036787253/2020\|EPI_ISL_728368\| | ···················· | ···················· | ····················· | ···················· | ···················· | ···················· |
| Y | 8542 hCoV-19/USA/MI-UM-10036818192/2020\|EPI_ISL_728390\| | ···················· | ···················· | ····················· | ···················· | ···················· | ···················· |
| Y | 8544 hCoV-19/USA/MI-UM-10036803164/2020\|EPI_ISL_728392\| | ···················· | ···················· | ····················· | ···················· | ···················· | ···················· |
| Y | 8545 hCoV-19/USA/MI-UM-10036801056/2020\|EPI_ISL_728393\| | ···················· | ···················· | ····················· | ···················· | ···················· | ···················· |
| Y | 8546 hCoV-19/USA/MI-UM-10036806044/2020\|EPI_ISL_728394\| | ···················· | ···················· | ····················· | ···················· | ···················· | ···················· |
| Y | 8547 hCoV-19/USA/MI-UM-10036804974/2020\|EPI_ISL_728395\| | ···················· | ···················· | ····················· | ···················· | ···················· | ···················· |
| Y | 8548 hCoV-19/USA/MI-UM-10036819118/2020\|EPI_ISL_728396\| | ···················· | ···················· | ····················· | ···················· | ···················· | ···················· |
| Y | 8549 hCoV-19/USA/MI-UM-10036813555/2020\|EPI_ISL_728397\| | ···················· | ···················· | ····················· | ···················· | ···················· | ···················· |
| Y | 8550 hCoV-19/USA/MI-UM-10036816187/2020\|EPI_ISL_728398\| | ···················· | ···················· | ····················· | ···················· | ···················· | ···················· |
| Y | 8551 hCoV-19/USA/MI-UM-10036821294/2020\|EPI_ISL_728399\| | ···················· | ···················· | ····················· | ···················· | ···················· | ···················· |
| Y | 8552 hCoV-19/Germany/DE-SH-ChVir20743/2020\|EPI_ISL_7296 | ···················· | ···················· | ····················· | ···················· | ···················· | ···················· |
| Y | 8553 hCoV-19/Germany/DE-SH-ChVir20744/2020\|EPI_ISL_7296 | ···················· | ···················· | ····················· | ···················· | ···················· | ···················· |

| | | | | | | | |
|---|---|---|---|---|---|---|---|
| Y | 8554 hCoV-19/Germany/DE-SH-ChVir20751/2020\|EPI_ISL_7296 | ···················· | ···················· | ···················· | ···················· | ···················· | ···················· |
| Y | 8555 hCoV-19/Germany/ChVir20769/2020\|EPI_ISL_729683\|202 | ···················· | ···················· | ···················· | ···················· | ···················· | ···················· |
| Y | 8556 hCoV-19/Germany/DE-SH-ChVir20758/2020\|EPI_ISL_7296 | ···················· | ···················· | ···················· | ···················· | ···················· | ···················· |
| Y | 8557 hCoV-19/Germany/DE-SH-ChVir20771/2020\|EPI_ISL_7296 | ···················· | ···················· | ···················· | ···················· | ···················· | ···················· |
| Y | 8558 hCoV-19/Germany/DE-SH-ChVir20775/2020\|EPI_ISL_7296 | ···················· | ···················· | ···················· | ···················· | ···················· | ···················· |
| Y | 8559 hCoV-19/Germany/DE-SH-ChVir20776/2020\|EPI_ISL_7296 | ···················· | ···················· | ···················· | ···················· | ···················· | ···················· |
| Y | 8560 hCoV-19/Germany/DE-SH-ChVir20777/2020\|EPI_ISL_7296 | ···················· | ···················· | ···················· | ···················· | ···················· | ···················· |
| Y | 8561 hCoV-19/Germany/DE-SH-ChVir20779/2020\|EPI_ISL_7296 | ···················· | ···················· | ···················· | ···················· | ···················· | ···················· |
| Y | 8562 hCoV-19/Germany/DE-SH-ChVir20784/2020\|EPI_ISL_7296 | ···················· | ···················· | ···················· | ···················· | ···················· | ···················· |
| Y | 8563 hCoV-19/Germany/DE-SH-ChVir20785/2020\|EPI_ISL_7296 | ···················· | ···················· | ···················· | ···················· | ···················· | ···················· |
| Y | 8564 hCoV-19/Germany/DE-SH-ChVir20786/2020\|EPI_ISL_7296 | ···················· | ···················· | ···················· | ···················· | ···················· | ···················· |
| Y | 8565 hCoV-19/Germany/DE-SH-ChVir20787/2020\|EPI_ISL_7296 | ···················· | ···················· | ···················· | ···················· | ···················· | ···················· |
| Y | 8566 hCoV-19/Germany/DE-SH-ChVir20789/2020\|EPI_ISL_7296 | ···················· | ···················· | ···················· | ···················· | ···················· | ···················· |
| Y | 8567 hCoV-19/Germany/DE-SH-ChVir20790/2020\|EPI_ISL_7296 | ···················· | ···················· | ···················· | ···················· | ···················· | ···················· |
| Y | 8568 hCoV-19/England/ALDP-C3D833/2020\|EPI_ISL_730667\|20 | ···················· | ···················· | ···················· | ···················· | ···················· | ···················· |
| Y | 8569 hCoV-19/England/ALDP-C3D815/2020\|EPI_ISL_730668\|20 | ···················· | ···················· | ···················· | ···················· | ···················· | ···················· |
| Y | 8570 hCoV-19/England/ALDP-C39DB0/2020\|EPI_ISL_730670\|20 | ···················· | ···················· | ···················· | ···················· | ···················· | ···················· |
| Y | 8571 hCoV-19/England/ALDP-C398EC/2020\|EPI_ISL_730671\|20 | ···················· | ···················· | ···················· | ···················· | ···················· | ···················· |
| Y | 8572 hCoV-19/England/ALDP-C3B6A4/2020\|EPI_ISL_730672\|20 | ···················· | ···················· | ···················· | ···················· | ···················· | ···················· |
| Y | 8573 hCoV-19/England/QEUH-C30FB3/2020\|EPI_ISL_730673\|20 | ···················· | ···················· | ···················· | ···················· | ···················· | ···················· |
| Y | 8574 hCoV-19/England/QEUH-C30D40/2020\|EPI_ISL_730676\|20 | ···················· | ···················· | ···················· | ···················· | ···················· | ···················· |
| Y | 8575 hCoV-19/England/ALDP-C3D8D9/2020\|EPI_ISL_730683\|20 | ···················· | ···················· | ···················· | ···················· | ···················· | ···················· |
| Y | 8576 hCoV-19/Wales/ALDP-C39934/2020\|EPI_ISL_730687\|2020 | ···················· | ···················· | ···················· | ···················· | ···················· | ···················· |
| Y | 8577 hCoV-19/Portugal/PT1764/2020\|EPI_ISL_731997\|2020-1 | ···················· | ···················· | ···················· | ···················· | ···················· | ···················· |
| Y | 8578 hCoV-19/Portugal/PT1766/2020\|EPI_ISL_731998\|2020-1 | ···················· | ···················· | ···················· | ···················· | ···················· | ···················· |
| Y | 8579 hCoV-19/Portugal/PT1768/2020\|EPI_ISL_731999\|2020-1 | ···················· | ···················· | ···················· | ···················· | ···················· | ···················· |
| Y | 8580 hCoV-19/USA/VA-DCLS-2455/2020\|EPI_ISL_734200\|2020- | ···················· | ···················· | ···················· | ···················· | ···················· | ···················· |
| Y | 8581 hCoV-19/USA/VA-DCLS-2456/2020\|EPI_ISL_734201\|2020- | ···················· | ···················· | ···················· | ···················· | ···················· | ···················· |
| Y | 8582 hCoV-19/USA/VA-DCLS-2457/2020\|EPI_ISL_734202\|2020- | ···················· | ···················· | ···················· | ···················· | ···················· | ···················· |
| Y | 8583 hCoV-19/USA/VA-DCLS-2459/2020\|EPI_ISL_734204\|2020- | ···················· | ···················· | ···················· | ···················· | ···················· | ···················· |
| Y | 8584 hCoV-19/USA/VA-DCLS-2460/2020\|EPI_ISL_734205\|2020- | ···················· | ···················· | ···················· | ···················· | ···················· | ···················· |
| Y | 8585 hCoV-19/USA/VA-DCLS-2462/2020\|EPI_ISL_734206\|2020- | ···················· | ···················· | ···················· | ···················· | ···················· | ···················· |
| Y | 8586 hCoV-19/USA/VA-DCLS-2217/2020\|EPI_ISL_734207\|2020- | ···················· | ···················· | ···················· | ···················· | ···················· | ···················· |
| Y | 8587 hCoV-19/USA/VA-DCLS-2222/2020\|EPI_ISL_734208\|2020- | ···················· | ···················· | ···················· | ···················· | ···················· | ···················· |
| Y | 8588 hCoV-19/USA/VA-DCLS-2225/2020\|EPI_ISL_734209\|2020- | ···················· | ···················· | ···················· | ···················· | ···················· | ···················· |
| Y | 8589 hCoV-19/USA/VA-DCLS-2227/2020\|EPI_ISL_734210\|2020- | ···················· | ···················· | ···················· | ···················· | ···················· | ···················· |
| Y | 8590 hCoV-19/USA/VA-DCLS-2230/2020\|EPI_ISL_734211\|2020- | ···················· | ···················· | ···················· | ···················· | ···················· | ···················· |
| Y | 8591 hCoV-19/USA/VA-DCLS-2231/2020\|EPI_ISL_734212\|2020- | ···················· | ···················· | ···················· | ···················· | ···················· | ···················· |
| Y | 8592 hCoV-19/USA/VA-DCLS-2232/2020\|EPI_ISL_734213\|2020- | ···················· | ···················· | ···················· | ···················· | ···················· | ···················· |
| Y | 8593 hCoV-19/USA/VA-DCLS-2234/2020\|EPI_ISL_734214\|2020- | ···················· | ·A·················· | ···················· | ···················· | ···················· | ···················· |
| Y | 8594 hCoV-19/USA/VA-DCLS-2235/2020\|EPI_ISL_734215\|2020- | ···················· | ···················· | ···················· | ···················· | ···················· | ···················· |
| Y | 8595 hCoV-19/USA/VA-DCLS-2236/2020\|EPI_ISL_734216\|2020- | ···················· | ···················· | ···················· | ···················· | ···················· | ···················· |
| Y | 8596 hCoV-19/USA/VA-DCLS-2237/2020\|EPI_ISL_734217\|2020- | ···················· | ···················· | ···················· | ···················· | ···················· | ···················· |
| Y | 8597 hCoV-19/USA/VA-DCLS-2238/2020\|EPI_ISL_734218\|2020- | ········T··········· | ·A·················· | ···················· | ···················· | ···················· | ···················· |
| Y | 8598 hCoV-19/USA/VA-DCLS-2239/2020\|EPI_ISL_734219\|2020- | ···················· | ···················· | ···················· | ···················· | ···················· | ···················· |
| Y | 8599 hCoV-19/USA/VA-DCLS-2240/2020\|EPI_ISL_734220\|2020- | ···················· | ···················· | ···················· | ···················· | ···················· | ···················· |
| Y | 8600 hCoV-19/USA/VA-DCLS-2241/2020\|EPI_ISL_734221\|2020- | ···················· | ···················· | ···················· | ···················· | ···················· | ···················· |
| Y | 8601 hCoV-19/USA/VA-DCLS-2243/2020\|EPI_ISL_734222\|2020- | ···················· | ·A·················· | ···················· | ···················· | ···················· | ···················· |
| Y | 8602 hCoV-19/USA/VA-DCLS-2245/2020\|EPI_ISL_734223\|2020- | ···················· | ······A············· | ···················· | ···················· | ···················· | ···················· |
| Y | 8603 hCoV-19/USA/VA-DCLS-2514/2020\|EPI_ISL_734224\|2020- | ···················· | ···················· | ···················· | ···················· | ···················· | ···················· |
| Y | 8604 hCoV-19/USA/VA-DCLS-2515/2020\|EPI_ISL_734225\|2020- | ···················· | ···················· | ···················· | ···················· | ···················· | ···················· |
| Y | 8605 hCoV-19/USA/VA-DCLS-2516/2020\|EPI_ISL_734226\|2020- | ···················· | ···················· | ···················· | ···················· | ···················· | ···················· |
| Y | 8606 hCoV-19/USA/VA-DCLS-2517/2020\|EPI_ISL_734227\|2020- | ···················· | ····A··············· | ···················· | ···················· | ···················· | ···················· |
| Y | 8607 hCoV-19/USA/VA-DCLS-2518/2020\|EPI_ISL_734228\|2020- | ···················· | ····A··············· | ···················· | ···················· | ···················· | ···················· |
| Y | 8608 hCoV-19/USA/VA-DCLS-2519/2020\|EPI_ISL_734229\|2020- | ···················· | ···················· | ···················· | ···················· | ···················· | ···················· |
| Y | 8609 hCoV-19/Bangladesh/CU-CTG-57/2020\|EPI_ISL_735500\|2 | ···················· | ···················· | ···················· | ···················· | ···················· | ···················· |
| Y | 8610 hCoV-19/Bangladesh/CU-CTG-71/2020\|EPI_ISL_735501\|2 | ···················· | ···················· | ···················· | ···················· | ···················· | ···················· |
| Y | 8611 hCoV-19/Bangladesh/CU-CTG-76/2020\|EPI_ISL_735502\|2 | ···················· | ···················· | ···················· | ···················· | ···················· | ···················· |
| Y | 8612 hCoV-19/Australia/ACT0094/2020\|EPI_ISL_735503\|2020 | ···················· | ···················· | ···················· | ···················· | ···················· | ···················· |
| Y | 8613 hCoV-19/Italy/LAZ-INMI-115/2020\|EPI_ISL_735510\|202 | ···················· | ···················· | ···················· | ···················· | ···················· | ···················· |
| Y | 8614 hCoV-19/England/MILK-C611D1/2020\|EPI_ISL_735512\|20 | ···················· | ···················· | ···················· | ···················· | ···················· | ···················· |
| Y | 8615 hCoV-19/England/MILK-C610C5/2020\|EPI_ISL_735515\|20 | ···················· | ···················· | ···················· | ···················· | ···················· | ···················· |
| Y | 8616 hCoV-19/England/MILK-C62AB7/2020\|EPI_ISL_735516\|20 | ···················· | ···················· | ···················· | ···················· | ···················· | ···················· |
| Y | 8617 hCoV-19/England/MILK-C63A6B/2020\|EPI_ISL_735528\|20 | ···················· | ···················· | ···················· | ···················· | ···················· | ···················· |
| Y | 8618 hCoV-19/England/MILK-C61353/2020\|EPI_ISL_735535\|20 | ···················· | ·A·················· | ···················· | ···················· | ···················· | ···A················ |
| Y | 8619 hCoV-19/England/MILK-C63A89/2020\|EPI_ISL_735537\|20 | ···················· | ···················· | ···················· | ···················· | ···················· | ···················· |
| Y | 8620 hCoV-19/England/MILK-C61DEC/2020\|EPI_ISL_735542\|20 | ···················· | ···················· | ···················· | ···················· | ···················· | ···················· |
| Y | 8621 hCoV-19/England/MILK-C61265/2020\|EPI_ISL_735543\|20 | ···················· | ···················· | ···················· | ···················· | ···················· | ···················· |
| Y | 8622 hCoV-19/England/MILK-C63209/2020\|EPI_ISL_735545\|20 | ···················· | ···················· | ···················· | ···················· | ···················· | ···················· |
| Y | 8623 hCoV-19/England/MILK-C61308/2020\|EPI_ISL_735548\|20 | ···················· | ···················· | ···················· | ···················· | ···················· | ···················· |
| Y | 8624 hCoV-19/England/MILK-C62AD5/2020\|EPI_ISL_735549\|20 | ···················· | ···················· | ···················· | ···················· | ···················· | ···················· |
| Y | 8625 hCoV-19/England/MILK-C61EDA/2020\|EPI_ISL_735551\|20 | ···················· | ···················· | ···················· | ···················· | ···················· | ···················· |
| Y | 8626 hCoV-19/England/MILK-C62B3C/2020\|EPI_ISL_735554\|20 | ···················· | ···················· | ···················· | ···················· | ···················· | ···················· |
| Y | 8627 hCoV-19/England/MILK-C61836/2020\|EPI_ISL_735555\|20 | ···················· | ···················· | ···················· | ···················· | ···················· | ···················· |
| Y | 8628 hCoV-19/Italy/CAM-TIGEM-13/2020\|EPI_ISL_736800\|202 | ···················· | ···················· | ···················· | ···················· | ···················· | ···················· |
| Y | 8629 hCoV-19/Italy/CAM-TIGEM-14/2020\|EPI_ISL_736801\|202 | ···················· | ·A·················· | ···················· | ···················· | ···················· | ···················· |
| Y | 8630 hCoV-19/Italy/CAM-TIGEM-15/2020\|EPI_ISL_736802\|202 | ···················· | ···················· | ···················· | ···················· | ···················· | ···················· |
| Y | 8631 hCoV-19/Italy/CAM-TIGEM-16/2020\|EPI_ISL_736803\|202 | ···················· | ·A·················· | ···················· | ···················· | ···················· | ···················· |
| Y | 8632 hCoV-19/Italy/CAM-TIGEM-22/2020\|EPI_ISL_736804\|202 | ···················· | ···················· | ···················· | ···················· | ······T············· | ···················· |
| Y | 8633 hCoV-19/Italy/CAM-TIGEM-26/2020\|EPI_ISL_736805\|202 | ···················· | ···················· | ···················· | ···················· | ···················· | ···················· |

| | | | | | | | |
|---|---|---|---|---|---|---|---|
| Y | 8634 hCoV-19/Italy/CAM-TIGEM-28/2020\|EPI_ISL_736806\|202 | ···················· | ···················· | ···················· | ···················· | ·····T·············· | ···················· |
| Y | 8635 hCoV-19/Italy/CAM-TIGEM-31/2020\|EPI_ISL_736807\|202 | ···················· | ···················· | ···················· | ···················· | ·····T·············· | ···················· |
| Y | 8636 hCoV-19/Italy/CAM-TIGEM-32/2020\|EPI_ISL_736808\|202 | ···················· | ···················· | ···················· | ···················· | ·····T·············· | ···················· |
| Y | 8637 hCoV-19/Italy/CAM-TIGEM-33/2020\|EPI_ISL_736809\|202 | ···················· | ·A·················· | ···················· | ···················· | ···················· | ···················· |
| Y | 8638 hCoV-19/Italy/CAM-TIGEM-34/2020\|EPI_ISL_736810\|202 | ···················· | ···················· | ···················· | ···················· | ·····T·············· | ···················· |
| Y | 8639 hCoV-19/Italy/CAM-TIGEM-35/2020\|EPI_ISL_736811\|202 | ···················· | ···················· | ···················· | ···················· | ···················· | ···················· |
| Y | 8640 hCoV-19/Italy/CAM-TIGEM-36/2020\|EPI_ISL_736812\|202 | ···················· | ···················· | ···················· | ···················· | ···················· | ···················· |
| Y | 8641 hCoV-19/Italy/CAM-TIGEM-37/2020\|EPI_ISL_736813\|202 | ···················· | ···················· | ···················· | ···················· | ···················· | ···················· |
| Y | 8642 hCoV-19/Italy/CAM-TIGEM-38/2020\|EPI_ISL_736814\|202 | ···················· | ···················· | ···················· | ···················· | ···················· | ···················· |
| Y | 8643 hCoV-19/Italy/CAM-TIGEM-39/2020\|EPI_ISL_736815\|202 | ···················· | ···················· | ···················· | ···················· | ···················· | ···················· |
| Y | 8644 hCoV-19/Italy/CAM-TIGEM-41/2020\|EPI_ISL_736816\|202 | ···················· | ···················· | ···················· | ···················· | ···················· | ···················· |
| Y | 8645 hCoV-19/Italy/CAM-TIGEM-42/2020\|EPI_ISL_736817\|202 | ···················· | ···················· | ···················· | ···················· | ···················· | ···················· |
| Y | 8646 hCoV-19/Italy/CAM-TIGEM-45/2020\|EPI_ISL_736818\|202 | ···················· | ···················· | ···················· | ···················· | ·····T·············· | ···················· |
| Y | 8647 hCoV-19/Italy/CAM-TIGEM-47/2020\|EPI_ISL_736819\|202 | ···················· | ···················· | ···················· | ···················· | ···················· | ···················· |
| Y | 8648 hCoV-19/Italy/CAM-TIGEM-48/2020\|EPI_ISL_736820\|202 | ···················· | ···················· | ···················· | ···················· | ·····T·············· | ···················· |
| Y | 8649 hCoV-19/Italy/CAM-TIGEM-49/2020\|EPI_ISL_736821\|202 | ···················· | ···················· | ···················· | ···················· | ···················· | ···················· |
| Y | 8650 hCoV-19/Italy/CAM-TIGEM-50/2020\|EPI_ISL_736822\|202 | ···················· | ···················· | ···················· | ···················· | ·····T·············· | ···················· |
| Y | 8651 hCoV-19/Italy/CAM-TIGEM-53/2020\|EPI_ISL_736823\|202 | ···················· | ···················· | ···················· | ···················· | ·····T·············· | ···················· |
| Y | 8652 hCoV-19/Italy/CAM-TIGEM-54/2020\|EPI_ISL_736824\|202 | ···················· | ···················· | ···················· | ···················· | ···················· | ···················· |
| Y | 8653 hCoV-19/Italy/CAM-TIGEM-57/2020\|EPI_ISL_736825\|202 | ···················· | ···················· | ···················· | ···················· | ···················· | ···················· |
| Y | 8654 hCoV-19/Italy/CAM-TIGEM-58/2020\|EPI_ISL_736826\|202 | ···················· | ···················· | ···················· | ···················· | ···················· | ···················· |
| Y | 8655 hCoV-19/Italy/CAM-TIGEM-59/2020\|EPI_ISL_736827\|202 | ···················· | ···················· | ···················· | ···················· | ·····T·············· | ···················· |
| Y | 8656 hCoV-19/Italy/CAM-TIGEM-60/2020\|EPI_ISL_736828\|202 | ···················· | ···················· | ···················· | ···················· | ·····T·············· | ···················· |
| Y | 8657 hCoV-19/Italy/CAM-TIGEM-61/2020\|EPI_ISL_736829\|202 | ···················· | ···················· | ···················· | ···················· | ···················· | ···················· |
| Y | 8658 hCoV-19/Italy/CAM-TIGEM-62/2020\|EPI_ISL_736830\|202 | ···················· | ···················· | ···················· | ···················· | ···················· | ···················· |
| Y | 8659 hCoV-19/Italy/CAM-TIGEM-63/2020\|EPI_ISL_736831\|202 | ···················· | ···················· | ···················· | ···················· | ···················· | ···················· |
| Y | 8660 hCoV-19/Italy/CAM-TIGEM-64/2020\|EPI_ISL_736832\|202 | ···················· | ···················· | ···················· | ···················· | ·····T·············· | ···················· |
| Y | 8661 hCoV-19/Italy/CAM-TIGEM-70/2020\|EPI_ISL_736833\|202 | ···················· | ···················· | ···················· | ···················· | ·····T·············· | ···················· |
| Y | 8662 hCoV-19/Italy/CAM-TIGEM-71/2020\|EPI_ISL_736834\|202 | ···················· | ···················· | ···················· | ···················· | ···················· | ···················· |
| Y | 8663 hCoV-19/Italy/CAM-TIGEM-74/2020\|EPI_ISL_736835\|202 | ·······T············ | ···················· | ···················· | ···················· | ···················· | ···················· |
| Y | 8664 hCoV-19/Italy/CAM-TIGEM-76/2020\|EPI_ISL_736836\|202 | ···················· | ···················· | ···················· | ···················· | ·····T·············· | ···················· |
| Y | 8665 hCoV-19/Italy/CAM-TIGEM-80/2020\|EPI_ISL_736837\|202 | ···················· | ·A·················· | ···················· | ···················· | ···················· | ···················· |
| Y | 8666 hCoV-19/Italy/CAM-TIGEM-88/2020\|EPI_ISL_736838\|202 | ···················· | ···················· | ···················· | ···················· | ···················· | ···················· |
| Y | 8667 hCoV-19/Italy/CAM-TIGEM-89/2020\|EPI_ISL_736839\|202 | ···················· | ···················· | ···················· | ···················· | ·····T·············· | ···················· |
| Y | 8668 hCoV-19/Italy/CAM-TIGEM-94/2020\|EPI_ISL_736840\|202 | ···················· | ···················· | ···················· | ···················· | ···················· | ···················· |
| Y | 8669 hCoV-19/Italy/CAM-TIGEM-95/2020\|EPI_ISL_736841\|202 | ···················· | ···················· | ···················· | ···················· | ···················· | ···················· |
| Y | 8670 hCoV-19/Italy/CAM-TIGEM-97/2020\|EPI_ISL_736842\|202 | ···················· | ···················· | ···················· | ···················· | ·····T·············· | ···················· |
| Y | 8671 hCoV-19/Italy/CAM-TIGEM-99/2020\|EPI_ISL_736843\|202 | ···················· | ···················· | ···················· | ···················· | ·····T·············· | ···················· |
| Y | 8672 hCoV-19/Italy/CAM-TIGEM-100/2020\|EPI_ISL_736844\|20 | ···················· | ···················· | ···················· | ···················· | ···················· | ···················· |
| Y | 8673 hCoV-19/Italy/CAM-TIGEM-102/2020\|EPI_ISL_736845\|20 | ···················· | ···A················ | ···················· | ···················· | ···················· | ···················· |
| Y | 8674 hCoV-19/Italy/CAM-TIGEM-105/2020\|EPI_ISL_736846\|20 | ···················· | ···A················ | ···················· | ···················· | ···················· | ···················· |
| Y | 8675 hCoV-19/Italy/CAM-TIGEM-107/2020\|EPI_ISL_736847\|20 | ···················· | ···················· | ···················· | ···················· | ·····T·············· | ···················· |
| Y | 8676 hCoV-19/Italy/CAM-TIGEM-126/2020\|EPI_ISL_736859\|20 | ···················· | ···················· | ···················· | ···················· | ·····T·············· | ···················· |
| Y | 8677 hCoV-19/Italy/CAM-TIGEM-132/2020\|EPI_ISL_736862\|20 | ···················· | ···················· | ···················· | ···················· | ···················· | ···················· |
| Y | 8678 hCoV-19/Italy/CAM-TIGEM-133/2020\|EPI_ISL_736863\|20 | ···················· | ···················· | ···················· | ···················· | ···················· | ···················· |
| Y | 8679 hCoV-19/Italy/CAM-TIGEM-135/2020\|EPI_ISL_736864\|20 | ···················· | ···················· | ···················· | ···················· | ···················· | ···················· |
| Y | 8680 hCoV-19/Italy/CAM-TIGEM-136/2020\|EPI_ISL_736865\|20 | ···················· | ···················· | ···················· | ···················· | ···················· | ···················· |
| Y | 8681 hCoV-19/Italy/CAM-TIGEM-138/2020\|EPI_ISL_736866\|20 | ···················· | ···················· | ···················· | ···················· | ···················· | ···················· |
| Y | 8682 hCoV-19/Italy/CAM-TIGEM-140/2020\|EPI_ISL_736867\|20 | ···················· | ···················· | ···················· | ···················· | ···················· | ···················· |
| Y | 8683 hCoV-19/Italy/CAM-TIGEM-141/2020\|EPI_ISL_736868\|20 | ···················· | ···················· | ···················· | ···················· | ···················· | ···················· |
| Y | 8684 hCoV-19/Italy/CAM-TIGEM-142/2020\|EPI_ISL_736869\|20 | ···················· | ···················· | ···················· | ···················· | ·····T·············· | ···················· |
| Y | 8685 hCoV-19/Italy/CAM-TIGEM-144/2020\|EPI_ISL_736870\|20 | ···················· | ···················· | ···················· | ···················· | ·····T·············· | ···················· |
| Y | 8686 hCoV-19/Italy/CAM-TIGEM-147/2020\|EPI_ISL_736871\|20 | ···················· | ···················· | ···················· | ···················· | ···················· | ···················· |
| Y | 8687 hCoV-19/Italy/CAM-TIGEM-148/2020\|EPI_ISL_736872\|20 | ···················· | ···················· | ···················· | ···················· | ···················· | ···················· |
| Y | 8688 hCoV-19/Italy/CAM-TIGEM-149/2020\|EPI_ISL_736873\|20 | ···················· | ···················· | ···················· | ···················· | ···················· | ···················· |
| Y | 8689 hCoV-19/Italy/CAM-TIGEM-150/2020\|EPI_ISL_736874\|20 | ···················· | ···················· | ···················· | ···················· | ···················· | ···················· |
| Y | 8690 hCoV-19/Italy/CAM-TIGEM-151/2020\|EPI_ISL_736875\|20 | ···················· | ···················· | ···················· | ···················· | ···················· | ···················· |
| Y | 8691 hCoV-19/Italy/CAM-TIGEM-154/2020\|EPI_ISL_736876\|20 | ···················· | ···················· | ···················· | ···················· | ···················· | ···················· |
| Y | 8692 hCoV-19/Italy/CAM-TIGEM-155/2020\|EPI_ISL_736877\|20 | ···················· | ···················· | ···················· | ···················· | ···················· | ···················· |
| Y | 8693 hCoV-19/Italy/CAM-TIGEM-157/2020\|EPI_ISL_736878\|20 | ···················· | ···················· | ···················· | ···················· | ···················· | ···················· |
| Y | 8694 hCoV-19/Italy/CAM-TIGEM-158/2020\|EPI_ISL_736879\|20 | ···················· | ···················· | ···················· | ···················· | ···················· | ···················· |
| Y | 8695 hCoV-19/Italy/CAM-TIGEM-159/2020\|EPI_ISL_736880\|20 | ···················· | ···················· | ···················· | ···················· | ···················· | ···················· |
| Y | 8696 hCoV-19/Italy/CAM-TIGEM-160/2020\|EPI_ISL_736881\|20 | ···················· | ···················· | ···················· | ···················· | ···················· | ···················· |
| Y | 8697 hCoV-19/Italy/CAM-TIGEM-161/2020\|EPI_ISL_736882\|20 | ···················· | ···················· | ···················· | ···················· | ···················· | ···················· |
| Y | 8698 hCoV-19/Italy/CAM-TIGEM-162/2020\|EPI_ISL_736883\|20 | ···················· | ···················· | ···················· | ···················· | ·····T·············· | ···················· |
| Y | 8699 hCoV-19/Italy/CAM-TIGEM-163/2020\|EPI_ISL_736884\|20 | ···················· | ···················· | ···················· | ···················· | ···················· | ···················· |
| Y | 8700 hCoV-19/Italy/CAM-TIGEM-164/2020\|EPI_ISL_736885\|20 | ···················· | ···················· | ···················· | ···················· | ···················· | ···················· |
| Y | 8701 hCoV-19/Italy/CAM-TIGEM-168/2020\|EPI_ISL_736886\|20 | ···················· | ···················· | ···················· | ···················· | ···················· | ···················· |
| Y | 8702 hCoV-19/Italy/CAM-TIGEM-169/2020\|EPI_ISL_736887\|20 | ···················· | ···················· | ···················· | ···················· | ···················· | ···················· |
| Y | 8703 hCoV-19/Italy/CAM-TIGEM-170/2020\|EPI_ISL_736888\|20 | ···················· | ···················· | ···················· | ···················· | ···················· | ···················· |
| Y | 8704 hCoV-19/Italy/CAM-TIGEM-171/2020\|EPI_ISL_736889\|20 | ···················· | ···················· | ···················· | ···················· | ·····T·············· | ···················· |
| Y | 8705 hCoV-19/England/NORT-1B72302/2020\|EPI_ISL_724869\|2 | ···················· | ···················· | ···················· | ···················· | ···················· | ···················· |
| Y | 8706 hCoV-19/England/NORT-1B7245A/2020\|EPI_ISL_724880\|2 | ···················· | ···················· | ···················· | ···················· | ···················· | ···················· |
| Y | 8707 hCoV-19/England/NORT-1B72478/2020\|EPI_ISL_724882\|2 | ···················· | ···················· | ···················· | ···················· | ···················· | ···················· |
| Y | 8708 hCoV-19/England/NORT-1B72496/2020\|EPI_ISL_724883\|2 | ···················· | ···················· | ···················· | ···················· | ···················· | ···················· |
| Y | 8709 hCoV-19/England/NORT-1B72566/2020\|EPI_ISL_724887\|2 | ···················· | ···················· | ···················· | ···················· | ···················· | ···················· |
| Y | 8710 hCoV-19/England/NORT-1B725DF/2020\|EPI_ISL_724888\|2 | ···················· | ···················· | ···················· | ···················· | ···················· | ···················· |
| Y | 8711 hCoV-19/England/NORT-1B725EE/2020\|EPI_ISL_724889\|2 | ···················· | ···················· | ···················· | ···················· | ···················· | ···················· |
| Y | 8713 hCoV-19/Wales/PHWC-498607/2020\|EPI_ISL_727101\|2020 | ···················· | ···················· | ···················· | ···················· | ···················· | ···················· |
| Y | 8714 hCoV-19/Wales/PHWC-498634/2020\|EPI_ISL_727103\|2020 | ···················· | ···················· | ···················· | ···················· | ···················· | ···················· |

| | | | | | | | | |
|---|---|---|---|---|---|---|---|---|
| Y | 8718 | hCoV-19/Wales/PHWC-498670/2020\|EPI_ISL_727107\|2020 | ···················· | ···················· | ···················· | ···················· | ···················· | ···················· |
| Y | 8719 | hCoV-19/Wales/PHWC-49868F/2020\|EPI_ISL_727108\|2020 | ···················· | ···················· | ···················· | ···················· | ···················· | ···················· |
| Y | 8721 | hCoV-19/Wales/PHWC-49878C/2020\|EPI_ISL_727120\|2020 | ···················· | ···················· | ···················· | ···················· | ···················· | ···················· |
| Y | 8722 | hCoV-19/Wales/PHWC-4987AA/2020\|EPI_ISL_727121\|2020 | ···················· | ···················· | ···················· | ···················· | ···················· | ···················· |
| Y | 8724 | hCoV-19/Wales/PHWC-498801/2020\|EPI_ISL_727127\|2020 | ···················· | ···················· | ···················· | ···················· | ···················· | ···················· |
| Y | 8725 | hCoV-19/Wales/PHWC-49882F/2020\|EPI_ISL_727128\|2020 | ···················· | ···················· | ···················· | ···················· | ···················· | ···················· |
| Y | 8728 | hCoV-19/Wales/PHWC-4988C5/2020\|EPI_ISL_727138\|2020 | ···················· | ···················· | ···················· | ···················· | ···················· | ···················· |
| Y | 8730 | hCoV-19/USA/MI-UM-10036824774/2020\|EPI_ISL_728400\| | ···················· | ···················· | ···················· | ···················· | ···················· | ···················· |
| Y | 8731 | hCoV-19/USA/MI-UM-10036820862/2020\|EPI_ISL_728401\| | ···················· | ···················· | ···················· | ···················· | ···················· | ···················· |
| Y | 8732 | hCoV-19/USA/MI-UM-10036814344/2020\|EPI_ISL_728402\| | ···················· | ·············A······ | ···················· | ···················· | ···················· | ···················· |
| Y | 8733 | hCoV-19/USA/MI-UM-10036813433/2020\|EPI_ISL_728403\| | ···················· | ···················· | ···················· | ···················· | ···················· | ···················· |
| Y | 8734 | hCoV-19/USA/MI-UM-10036822002/2020\|EPI_ISL_728404\| | ···················· | ···················· | ···················· | ···················· | ···················· | ···················· |
| Y | 8735 | hCoV-19/USA/MI-UM-10036818034/2020\|EPI_ISL_728406\| | ···················· | ···················· | ···················· | ···················· | ···················· | ···················· |
| Y | 8736 | hCoV-19/USA/MI-UM-10036823312/2020\|EPI_ISL_728408\| | ···················· | ···················· | ···················· | ···················· | ···················· | ···················· |
| Y | 8738 | hCoV-19/USA/MI-UM-10036821475/2020\|EPI_ISL_728410\| | ···················· | ···················· | ···················· | ···················· | ···················· | ···················· |
| Y | 8742 | hCoV-19/USA/MI-UM-10036817918/2020\|EPI_ISL_728414\| | ···················· | ···················· | ···················· | ···················· | ···················· | ···················· |
| Y | 8743 | hCoV-19/USA/MI-UM-10036821152/2020\|EPI_ISL_728415\| | ···················· | ·············A······ | ···················· | ···················· | ···················· | ···················· |
| Y | 8750 | hCoV-19/USA/MI-UM-10036826453/2020\|EPI_ISL_728425\| | ···················· | ···················· | ···················· | ···················· | ···················· | ···················· |
| Y | 8752 | hCoV-19/USA/MI-UM-10036819514/2020\|EPI_ISL_728427\| | ···················· | ···················· | ···················· | ···················· | ···················· | ···················· |
| Y | 8753 | hCoV-19/USA/MI-UM-10036825820/2020\|EPI_ISL_728428\| | ···················· | ···················· | ···················· | ···················· | ···················· | ···················· |
| Y | 8754 | hCoV-19/USA/MI-UM-10036820826/2020\|EPI_ISL_728429\| | ···················· | ···················· | ···················· | ···················· | ···················· | ···················· |
| Y | 8755 | hCoV-19/USA/MI-UM-10036826472/2020\|EPI_ISL_728430\| | ···················· | ···················· | ···················· | ···················· | ···················· | ···················· |
| Y | 8757 | hCoV-19/USA/MI-UM-10036826852/2020\|EPI_ISL_728432\| | ···················· | ······A············· | ···················· | ···················· | ···················· | ···················· |
| Y | 8758 | hCoV-19/USA/MI-UM-10036817055/2020\|EPI_ISL_728433\| | ···················· | ···················· | ···················· | ···················· | ···················· | ···················· |
| Y | 8759 | hCoV-19/USA/MI-UM-10036824591/2020\|EPI_ISL_728436\| | ···················· | ···················· | ···················· | ···················· | ···················· | ···················· |
| Y | 8761 | hCoV-19/USA/MI-UM-10036834507/2020\|EPI_ISL_728438\| | ···················· | ···················· | ···················· | ···················· | ···················· | ···················· |
| Y | 8768 | hCoV-19/USA/MI-UM-10036848661/2020\|EPI_ISL_728455\| | ···················· | ···················· | ···················· | ···················· | ···················· | ···················· |
| Y | 8773 | hCoV-19/USA/MI-UM-10036832225/2020\|EPI_ISL_728462\| | ···················· | ···················· | ···················· | ···················· | ···················· | ···················· |
| Y | 8774 | hCoV-19/USA/MI-UM-10036835234/2020\|EPI_ISL_728463\| | ···················· | ···················· | ···················· | ···················· | ···················· | ···················· |
| Y | 8775 | hCoV-19/USA/MI-UM-10036835650/2020\|EPI_ISL_728464\| | ···················· | ···················· | ···················· | ···················· | ···················· | ···················· |
| Y | 8776 | hCoV-19/USA/MI-UM-10036835250/2020\|EPI_ISL_728465\| | ···················· | ···················· | ···················· | ···················· | ···················· | ···················· |
| Y | 8779 | hCoV-19/Germany/DE-SH-ChVir20792/2020\|EPI_ISL_7297 | ···················· | ···················· | ···················· | ···················· | ···················· | ···················· |
| Y | 8780 | hCoV-19/Germany/DE-SH-ChVir20793/2020\|EPI_ISL_7297 | ···················· | ···················· | ···················· | ···················· | ···················· | ···················· |
| Y | 8781 | hCoV-19/Germany/DE-SH-ChVir20794/2020\|EPI_ISL_7297 | ···················· | ···················· | ···················· | ···················· | ···················· | ···················· |
| Y | 8782 | hCoV-19/Germany/DE-SH-ChVir20795/2020\|EPI_ISL_7297 | ···················· | ···················· | ···················· | ···················· | ···················· | ···················· |
| Y | 8783 | hCoV-19/Germany/DE-SH-ChVir20796/2020\|EPI_ISL_7297 | ···················· | ···················· | ···················· | ···················· | ···················· | ···················· |
| Y | 8784 | hCoV-19/Germany/DE-SH-ChVir20797/2020\|EPI_ISL_7297 | ···················· | ···················· | ···················· | ···················· | ···················· | ···················· |
| Y | 8785 | hCoV-19/Germany/DE-SH-ChVir20798/2020\|EPI_ISL_7297 | ···················· | ···················· | ···················· | ···················· | ···················· | ···················· |
| Y | 8786 | hCoV-19/Germany/DE-SH-ChVir20800/2020\|EPI_ISL_7297 | ···················· | ···················· | ···················· | ···················· | ···················· | ···················· |
| Y | 8787 | hCoV-19/Germany/DE-SH-ChVir20801/2020\|EPI_ISL_7297 | ···················· | ···················· | ···················· | ···················· | ···················· | ···················· |
| Y | 8788 | hCoV-19/Germany/DE-SH-ChVir20803/2020\|EPI_ISL_7297 | ···················· | ·A·················· | ···················· | ···················· | ···················· | ···················· |
| Y | 8789 | hCoV-19/Germany/DE-SH-ChVir20807/2020\|EPI_ISL_7297 | ···················· | ···················· | ···················· | ···················· | ···················· | ···················· |
| Y | 8790 | hCoV-19/Germany/DE-SH-ChVir20809/2020\|EPI_ISL_7297 | ···················· | ···················· | ···················· | ···················· | ···················· | ···················· |
| Y | 8791 | hCoV-19/Germany/DE-SH-ChVir20811/2020\|EPI_ISL_7297 | ···················· | ···················· | ···················· | ···················· | ···················· | ···················· |
| Y | 8792 | hCoV-19/Germany/DE-SH-ChVir20812/2020\|EPI_ISL_7297 | ···················· | ···················· | ···················· | ···················· | ···················· | ···················· |
| Y | 8793 | hCoV-19/Germany/DE-SH-ChVir20815/2020\|EPI_ISL_7297 | ···················· | ···················· | ···················· | ···················· | ···················· | ···················· |
| Y | 8794 | hCoV-19/Germany/DE-SH-ChVir20832/2020\|EPI_ISL_7297 | ···················· | ···················· | ···················· | ···················· | ···················· | ···················· |
| Y | 8795 | hCoV-19/Germany/DE-SH-ChVir20818/2020\|EPI_ISL_7297 | ···················· | ···················· | ···················· | ···················· | ···················· | ···················· |
| Y | 8796 | hCoV-19/Germany/DE-SH-ChVir20819/2020\|EPI_ISL_7297 | ···················· | ···················· | ···················· | ···················· | ···················· | ···················· |
| Y | 8797 | hCoV-19/Germany/ChVir20820/2020\|EPI_ISL_729718\|202 | ···················· | ···················· | ···················· | ···················· | ···················· | ···················· |
| Y | 8798 | hCoV-19/Germany/DE-SH-ChVir20822/2020\|EPI_ISL_7297 | ···················· | ·A·················· | ···················· | ···················· | ···················· | ···················· |
| Y | 8799 | hCoV-19/Germany/DE-SH-ChVir20823/2020\|EPI_ISL_7297 | ···················· | ···················· | ···················· | ···················· | ···················· | ···················· |
| Y | 8800 | hCoV-19/Germany/DE-SH-ChVir20824/2020\|EPI_ISL_7297 | ···················· | ···················· | ···················· | ···················· | ···················· | ···················· |
| Y | 8801 | hCoV-19/Germany/DE-SH-ChVir20825/2020\|EPI_ISL_7297 | ···················· | ···················· | ···················· | ···················· | ···················· | ···················· |
| Y | 8802 | hCoV-19/Germany/DE-SH-ChVir20841/2020\|EPI_ISL_7297 | ···················· | ···················· | ···················· | ···················· | ···················· | ···················· |
| Y | 8803 | hCoV-19/Germany/DE-SH-ChVir20845/2020\|EPI_ISL_7297 | ···················· | ···················· | ···················· | ···················· | ···················· | ···················· |
| Y | 8804 | hCoV-19/Germany/DE-SH-ChVir20846/2020\|EPI_ISL_7297 | ···················· | ···················· | ···················· | ···················· | ···················· | ···················· |
| Y | 8805 | hCoV-19/Germany/DE-SH-ChVir20847/2020\|EPI_ISL_7297 | ···················· | ·A·················· | ···················· | ···················· | ···················· | ···················· |
| Y | 8806 | hCoV-19/Germany/DE-SH-ChVir20848/2020\|EPI_ISL_7297 | ···················· | ·A·················· | ···················· | ···················· | ···················· | ···················· |
| Y | 8807 | hCoV-19/Germany/DE-SH-ChVir20826/2020\|EPI_ISL_7297 | ···················· | ···················· | ···················· | ···················· | ···················· | ···················· |
| Y | 8808 | hCoV-19/Germany/DE-SH-ChVir20827/2020\|EPI_ISL_7297 | ···················· | ·A····A············· | ···················· | ···················· | ···················· | ···················· |
| Y | 8809 | hCoV-19/Germany/DE-SH-ChVir20828/2020\|EPI_ISL_7297 | ···················· | ···················· | ···················· | ···················· | ···················· | ···················· |
| Y | 8810 | hCoV-19/Germany/DE-SH-ChVir20829/2020\|EPI_ISL_7297 | ···················· | ···················· | ···················· | ···················· | ···················· | ···················· |
| Y | 8811 | hCoV-19/Germany/DE-SH-ChVir20833/2020\|EPI_ISL_7297 | ···················· | ···················· | ···················· | ···················· | ···················· | ···················· |
| Y | 8812 | hCoV-19/Germany/DE-SH-ChVir20835/2020\|EPI_ISL_7297 | ···················· | ···················· | ···················· | ···················· | ···················· | ···················· |
| Y | 8813 | hCoV-19/Germany/DE-SH-ChVir20836/2020\|EPI_ISL_7297 | ···················· | ···················· | ···················· | ···················· | ···················· | ···················· |
| Y | 8814 | hCoV-19/Brazil/RS-6195/2020\|EPI_ISL_729794\|2020-05 | ···················· | ···················· | ···················· | ···················· | ···················· | ···················· |
| Y | 8815 | hCoV-19/England/ALDP-C39E80/2020\|EPI_ISL_730700\|20 | ···················· | ···················· | ···················· | ···················· | ···················· | ···················· |
| Y | 8816 | hCoV-19/England/ALDP-C3A951/2020\|EPI_ISL_730701\|20 | ···················· | ···················· | ···················· | ···················· | ···················· | ···················· |
| Y | 8817 | hCoV-19/England/ALDP-C3D8BB/2020\|EPI_ISL_730704\|20 | ···················· | ···················· | ···················· | ···················· | ···················· | ···················· |
| Y | 8818 | hCoV-19/England/QEUH-C31996/2020\|EPI_ISL_730705\|20 | ···················· | ···················· | ···················· | ···················· | ···················· | ···············A···· |
| Y | 8819 | hCoV-19/England/ALDP-C3AA03/2020\|EPI_ISL_730707\|20 | ···················· | ···················· | ···················· | ···················· | ···················· | ···················· |
| Y | 8820 | hCoV-19/England/ALDP-C39FE7/2020\|EPI_ISL_730708\|20 | ···················· | ···················· | ···················· | ···················· | ···················· | ···················· |
| Y | 8821 | hCoV-19/England/QEUH-C314FF/2020\|EPI_ISL_730710\|20 | ···················· | ···················· | ···················· | ···················· | ···················· | ···················· |
| Y | 8822 | hCoV-19/England/ALDP-C3B1FE/2020\|EPI_ISL_730713\|20 | ···················· | ···················· | ···················· | ···················· | ···················· | ···················· |
| Y | 8823 | hCoV-19/England/ALDP-C3B219/2020\|EPI_ISL_730714\|20 | ···················· | ···················· | ···················· | ···················· | ···················· | ···················· |
| Y | 8824 | hCoV-19/England/ALDP-C39B5C/2020\|EPI_ISL_730717\|20 | ···················· | ···················· | ···················· | ···················· | ···················· | ···················· |
| Y | 8825 | hCoV-19/England/QEUH-C30F95/2020\|EPI_ISL_730718\|20 | ···················· | ···················· | ···················· | ···················· | ···················· | ···················· |
| Y | 8826 | hCoV-19/England/ALDP-C3A13B/2020\|EPI_ISL_730721\|20 | ···················· | ···················· | ···················· | ···················· | ···················· | ···················· |
| Y | 8827 | hCoV-19/England/ALDP-C3DD70/2020\|EPI_ISL_730722\|20 | ···················· | ·A·················· | ···················· | ···················· | ···················· | ···A················ |

| | | | | | | | | |
|---|---|---|---|---|---|---|---|---|
| Y | 8828 | hCoV-19/England/QEUH-C314E0/2020\|EPI_ISL_730723\|20 | ···················· | ···················· | ···················· | ···················· | ······T············· | ···················· |
| Y | 8829 | hCoV-19/England/ALDP-C3AE25/2020\|EPI_ISL_730724\|20 | ···················· | ···················· | ···················· | ···················· | ···················· | ···················· |
| Y | 8830 | hCoV-19/England/ALDP-C39873/2020\|EPI_ISL_730725\|20 | ···················· | ···················· | ···················· | ···················· | ···················· | ···················· |
| Y | 8831 | hCoV-19/England/ALDP-C3B695/2020\|EPI_ISL_730726\|20 | ···················· | ···················· | ···················· | ···················· | ···················· | ···················· |
| Y | 8832 | hCoV-19/England/ALDP-C3D7EB/2020\|EPI_ISL_730729\|20 | ···················· | ···················· | ···················· | ···················· | ···················· | ···················· |
| Y | 8833 | hCoV-19/England/ALDP-C3A9D9/2020\|EPI_ISL_730730\|20 | ···················· | ···················· | ···················· | ···················· | ···················· | ···················· |
| Y | 8834 | hCoV-19/England/ALDP-C3DBB2/2020\|EPI_ISL_730731\|20 | ···················· | ···················· | ···················· | ···················· | ···················· | ···················· |
| Y | 8835 | hCoV-19/England/ALDP-C3DDDA/2020\|EPI_ISL_730732\|20 | ···················· | ···················· | ···················· | ···················· | ···················· | ···················· |
| Y | 8836 | hCoV-19/England/ALDP-C3A05C/2020\|EPI_ISL_730736\|20 | ···················· | ···················· | ···················· | ···················· | ···················· | ···················· |
| Y | 8837 | hCoV-19/England/QEUH-C317C9/2020\|EPI_ISL_730737\|20 | ···················· | ···················· | ···················· | ···················· | ···················· | ···················· |
| Y | 8838 | hCoV-19/England/ALDP-C3B677/2020\|EPI_ISL_730738\|20 | ···················· | ···················· | ···················· | ···················· | ···················· | ···················· |
| Y | 8839 | hCoV-19/England/ALDP-C3A960/2020\|EPI_ISL_730739\|20 | ···················· | ···················· | ···················· | ···················· | ···················· | ···················· |
| Y | 8840 | hCoV-19/England/ALDP-C3B1A3/2020\|EPI_ISL_730741\|20 | ···················· | ···················· | ···················· | ···················· | ···················· | ···················· |
| Y | 8841 | hCoV-19/England/ALDP-C3DDCB/2020\|EPI_ISL_730754\|20 | ···················· | ···················· | ···················· | ···················· | ···················· | ···················· |
| Y | 8842 | hCoV-19/England/ALDP-C39B4D/2020\|EPI_ISL_730755\|20 | ···················· | ···················· | ···················· | ···················· | ···················· | ···················· |
| Y | 8843 | hCoV-19/England/ALDP-C3B686/2020\|EPI_ISL_730756\|20 | ···················· | ···················· | ···················· | ···················· | ···················· | ···················· |
| Y | 8844 | hCoV-19/England/QEUH-C30E10/2020\|EPI_ISL_730765\|20 | ···················· | ···················· | ···················· | ···················· | ···················· | ···················· |
| Y | 8845 | hCoV-19/England/ALDP-C39E71/2020\|EPI_ISL_730766\|20 | ···················· | ···················· | ···················· | ···················· | ···················· | ···················· |
| Y | 8846 | hCoV-19/England/ALDP-C39B10/2020\|EPI_ISL_730767\|20 | ···················· | ···················· | ···················· | ···················· | ····T··············· | ···················· |
| Y | 8847 | hCoV-19/England/ALDP-C3B6B3/2020\|EPI_ISL_730770\|20 | ···················· | ···················· | ···················· | ···················· | ···················· | ···A················ |
| Y | 8848 | hCoV-19/England/ALDP-C3AEDA/2020\|EPI_ISL_730772\|20 | ···················· | ···················· | ···················· | ···················· | ···················· | ···A················ |
| Y | 8849 | hCoV-19/England/ALDP-C3D5C3/2020\|EPI_ISL_730773\|20 | ···················· | ···················· | ···················· | ···················· | ···················· | ···················· |
| Y | 8850 | hCoV-19/England/ALDP-C3D806/2020\|EPI_ISL_730774\|20 | ···················· | ···················· | ···················· | ···················· | ···················· | ···················· |
| Y | 8851 | hCoV-19/England/ALDP-C3B6C2/2020\|EPI_ISL_730775\|20 | ···················· | ···················· | ···················· | ···················· | ···················· | ···················· |
| Y | 8852 | hCoV-19/England/ALDP-C3B52F/2020\|EPI_ISL_730776\|20 | ···················· | ···················· | ···················· | ···················· | ···················· | ···················· |
| Y | 8853 | hCoV-19/England/QEUH-C319A5/2020\|EPI_ISL_730777\|20 | ···················· | ···················· | ···················· | ···················· | ···················· | ···················· |
| Y | 8854 | hCoV-19/England/ALDP-C3A195/2020\|EPI_ISL_730779\|20 | ···················· | ···················· | ···················· | ···················· | ···················· | ···················· |
| Y | 8855 | hCoV-19/England/QEUH-C319E1/2020\|EPI_ISL_730780\|20 | ···················· | ···················· | ···················· | ···················· | ···················· | ···················· |
| Y | 8856 | hCoV-19/England/ALDP-C39B3E/2020\|EPI_ISL_730783\|20 | ···················· | ···················· | ···················· | ···················· | ···················· | ···················· |
| Y | 8857 | hCoV-19/England/QEUH-C31477/2020\|EPI_ISL_730785\|20 | ···················· | ···················· | ···················· | ···················· | ···················· | ···················· |
| Y | 8858 | hCoV-19/USA/VA-DCLS-2458/2020\|EPI_ISL_734203\|2020- | ···················· | ···················· | ···················· | ···················· | ···················· | ···················· |
| Y | 8859 | hCoV-19/USA/VA-DCLS-2520/2020\|EPI_ISL_734230\|2020- | ···················· | ···················· | ···················· | ···················· | ···················· | ···················· |
| Y | 8860 | hCoV-19/USA/VA-DCLS-2522/2020\|EPI_ISL_734231\|2020- | ···················· | ···················· | ···················· | ···················· | ···················· | ···················· |
| Y | 8861 | hCoV-19/USA/VA-DCLS-2162/2020\|EPI_ISL_734232\|2020- | ···················· | ···················· | ···················· | ···················· | ···················· | ···················· |
| Y | 8862 | hCoV-19/USA/VA-DCLS-2145/2020\|EPI_ISL_734233\|2020- | ···················· | ·A·················· | ···················· | ···················· | ···················· | ···················· |
| Y | 8863 | hCoV-19/USA/VA-DCLS-2147/2020\|EPI_ISL_734234\|2020- | ···················· | ···················· | ···················· | ···················· | ···················· | ···················· |
| Y | 8864 | hCoV-19/USA/VA-DCLS-2149/2020\|EPI_ISL_734235\|2020- | ···················· | ···················· | ···················· | ···················· | ···················· | ···················· |
| Y | 8865 | hCoV-19/USA/VA-DCLS-2153/2020\|EPI_ISL_734236\|2020- | ···················· | ···················· | ···················· | ···················· | ···················· | ···················· |
| Y | 8866 | hCoV-19/USA/VA-DCLS-2154/2020\|EPI_ISL_734237\|2020- | ···················· | ···················· | ···················· | ···················· | ···················· | ···················· |
| Y | 8867 | hCoV-19/USA/VA-DCLS-2155/2020\|EPI_ISL_734238\|2020- | ···················· | ···················· | ···················· | ···················· | ···················· | ···················· |
| Y | 8868 | hCoV-19/USA/VA-DCLS-2157/2020\|EPI_ISL_734239\|2020- | ···················· | ···················· | ···················· | ···················· | ···················· | ···················· |
| Y | 8869 | hCoV-19/USA/VA-DCLS-2158/2020\|EPI_ISL_734240\|2020- | ···················· | ···················· | ···················· | ···················· | ···················· | ···················· |
| Y | 8870 | hCoV-19/USA/VA-DCLS-2159/2020\|EPI_ISL_734241\|2020- | ···················· | ···················· | ···················· | ···················· | ···················· | ···················· |
| Y | 8871 | hCoV-19/USA/VA-DCLS-2160/2020\|EPI_ISL_734242\|2020- | ···················· | ···················· | ···················· | ···················· | ···················· | ···················· |
| Y | 8872 | hCoV-19/USA/VA-DCLS-2161/2020\|EPI_ISL_734243\|2020- | ···················· | ···················· | ···················· | ···················· | ···················· | ···················· |
| Y | 8873 | hCoV-19/USA/VA-DCLS-2163/2020\|EPI_ISL_734244\|2020- | ···················· | ···················· | ···················· | ···················· | ···················· | ···················· |
| Y | 8874 | hCoV-19/USA/VA-DCLS-2164/2020\|EPI_ISL_734245\|2020- | ···················· | ·A·················· | ···················· | ···················· | ···················· | ···················· |
| Y | 8875 | hCoV-19/USA/VA-DCLS-2193/2020\|EPI_ISL_734246\|2020- | ···················· | ·A·················· | ···················· | ···················· | ···················· | ···················· |
| Y | 8876 | hCoV-19/USA/VA-DCLS-2194/2020\|EPI_ISL_734247\|2020- | ···················· | ·A·················· | ···················· | ···················· | ···················· | ···················· |
| Y | 8877 | hCoV-19/USA/VA-DCLS-2195/2020\|EPI_ISL_734248\|2020- | ···················· | ·A·················· | ···················· | ···················· | ···················· | ···················· |
| Y | 8878 | hCoV-19/USA/VA-DCLS-2196/2020\|EPI_ISL_734249\|2020- | ···················· | ·A·················· | ···················· | ···················· | ···················· | ···················· |
| Y | 8879 | hCoV-19/USA/VA-DCLS-2197/2020\|EPI_ISL_734250\|2020- | ···················· | ·A·················· | ···················· | ···················· | ···················· | ···················· |
| Y | 8880 | hCoV-19/USA/VA-DCLS-2198/2020\|EPI_ISL_734251\|2020- | ···················· | ·A·················· | ···················· | ···················· | ···················· | ···················· |
| Y | 8881 | hCoV-19/USA/VA-DCLS-2199/2020\|EPI_ISL_734252\|2020- | ···················· | ·A·················· | ···················· | ···················· | ···················· | ···················· |
| Y | 8882 | hCoV-19/USA/VA-DCLS-2537/2020\|EPI_ISL_734253\|2020- | ···················· | ···················· | ···················· | ···················· | ·········T·········· | ···················· |
| Y | 8883 | hCoV-19/USA/VA-DCLS-2538/2020\|EPI_ISL_734254\|2020- | ···················· | ···················· | ···················· | ···················· | ···················· | ···················· |
| Y | 8884 | hCoV-19/USA/VA-DCLS-2320/2020\|EPI_ISL_734255\|2020- | ···················· | ···················· | ···················· | ···················· | ···················· | ···················· |
| Y | 8885 | hCoV-19/USA/VA-DCLS-2512/2020\|EPI_ISL_734256\|2020- | ···················· | ···················· | ···················· | ···················· | ···················· | ···················· |
| Y | 8886 | hCoV-19/USA/VA-DCLS-2297/2020\|EPI_ISL_734257\|2020- | ···················· | ···················· | ···················· | ···················· | ···················· | ···················· |
| Y | 8887 | hCoV-19/USA/VA-DCLS-2300/2020\|EPI_ISL_734258\|2020- | ···················· | ···················· | ···················· | ···················· | ···················· | ···················· |
| Y | 8888 | hCoV-19/USA/VA-DCLS-2301/2020\|EPI_ISL_734259\|2020- | ···················· | ···················· | ···················· | ···················· | ···················· | ···················· |
| Y | 8889 | hCoV-19/USA/VA-DCLS-2304/2020\|EPI_ISL_734260\|2020- | ···················· | ···················· | ···················· | ···················· | ···················· | ···················· |
| Y | 8890 | hCoV-19/USA/VA-DCLS-2305/2020\|EPI_ISL_734261\|2020- | ···················· | ···················· | ···················· | ···················· | ···················· | ···················· |
| Y | 8891 | hCoV-19/USA/VA-DCLS-2307/2020\|EPI_ISL_734262\|2020- | ···················· | ···················· | ···················· | ···················· | ···················· | ···················· |
| Y | 8892 | hCoV-19/USA/VA-DCLS-2309/2020\|EPI_ISL_734263\|2020- | ···················· | ···················· | ···················· | ···················· | ···················· | ···················· |
| Y | 8893 | hCoV-19/USA/VA-DCLS-2311/2020\|EPI_ISL_734264\|2020- | ···················· | ···················· | ···················· | ···················· | ···················· | ···················· |
| Y | 8894 | hCoV-19/USA/VA-DCLS-2314/2020\|EPI_ISL_734265\|2020- | ···················· | ·A·················· | ···················· | ···················· | ···················· | ···················· |
| Y | 8895 | hCoV-19/USA/VA-DCLS-2316/2020\|EPI_ISL_734266\|2020- | ···················· | ·A·················· | ···················· | ···················· | ···················· | ···················· |
| Y | 8896 | hCoV-19/USA/VA-DCLS-2317/2020\|EPI_ISL_734267\|2020- | ···················· | ···················· | ···················· | ···················· | ···················· | ···················· |
| Y | 8897 | hCoV-19/USA/VA-DCLS-2318/2020\|EPI_ISL_734268\|2020- | ···················· | ·A·················· | ···················· | ···················· | ···················· | ···················· |
| Y | 8898 | hCoV-19/USA/VA-DCLS-2319/2020\|EPI_ISL_734269\|2020- | ···················· | ···················· | ···················· | ···················· | ···················· | ···················· |
| Y | 8899 | hCoV-19/USA/VA-DCLS-2322/2020\|EPI_ISL_734270\|2020- | ···················· | ···················· | ···················· | ···················· | ···················· | ···················· |
| Y | 8900 | hCoV-19/USA/VA-DCLS-2323/2020\|EPI_ISL_734271\|2020- | ···················· | ·A·················· | ···················· | ···················· | ···················· | ···················· |
| Y | 8901 | hCoV-19/USA/VA-DCLS-2324/2020\|EPI_ISL_734272\|2020- | ···················· | ···················· | ···················· | ···················· | ·········T·········· | ···················· |
| Y | 8902 | hCoV-19/USA/VA-DCLS-2325/2020\|EPI_ISL_734273\|2020- | ···················· | ·A·················· | ···················· | ···················· | ···················· | ···················· |
| Y | 8903 | hCoV-19/USA/VA-DCLS-2326/2020\|EPI_ISL_734274\|2020- | ···················· | ·A·················· | ···················· | ···················· | ···················· | ···················· |
| Y | 8904 | hCoV-19/USA/VA-DCLS-2328/2020\|EPI_ISL_734275\|2020- | ···················· | ·A·················· | ···················· | ···················· | ···················· | ···················· |
| Y | 8905 | hCoV-19/USA/VA-DCLS-2329/2020\|EPI_ISL_734276\|2020- | ···················· | ·A·················· | ···················· | ···················· | ···················· | ···················· |
| Y | 8906 | hCoV-19/USA/VA-DCLS-2330/2020\|EPI_ISL_734277\|2020- | ···················· | ···················· | ···················· | ···················· | ···················· | ···················· |
| Y | 8907 | hCoV-19/USA/VA-DCLS-2332/2020\|EPI_ISL_734278\|2020- | ···················· | ···················· | ···················· | ···················· | ···················· | ···················· |

| | | | | | | | |
|---|---|---|---|---|---|---|---|
| Y | 8908 hCoV-19/USA/VA-DCLS-2333/2020\|EPI_ISL_734279\|2020- | ···················· | ·A·················· | ···················· | ···················· | ···················· | ···················· |
| Y | 8909 hCoV-19/USA/VA-DCLS-2503/2020\|EPI_ISL_734280\|2020- | ···················· | ···A················ | ···················· | ···················· | ···················· | ···················· |
| Y | 8910 hCoV-19/USA/VA-DCLS-2505/2020\|EPI_ISL_734281\|2020- | ···················· | ···················· | ···················· | ···················· | ···················· | ···················· |
| Y | 8911 hCoV-19/USA/VA-DCLS-2507/2020\|EPI_ISL_734282\|2020- | ···················· | ···················· | ···················· | ···················· | ···················· | ···················· |
| Y | 8912 hCoV-19/USA/VA-DCLS-2508/2020\|EPI_ISL_734283\|2020- | ···················· | ···················· | ···················· | ···················· | ···················· | ···················· |
| Y | 8913 hCoV-19/USA/VA-DCLS-2509/2020\|EPI_ISL_734284\|2020- | ···················· | ···················· | ···················· | ···················· | ···················· | ···················· |
| Y | 8914 hCoV-19/USA/VA-DCLS-2510/2020\|EPI_ISL_734285\|2020- | ···················· | ···················· | ···················· | ···················· | ···················· | ···················· |
| Y | 8915 hCoV-19/USA/VA-DCLS-2511/2020\|EPI_ISL_734286\|2020- | ···················· | ···················· | ···················· | ···················· | ···················· | ···················· |
| Y | 8916 hCoV-19/USA/NY-Wadsworth-250933-01/2020\|EPI_ISL_73 | ···················· | ···················· | ···················· | ···················· | ···················· | ···················· |
| Y | 8917 hCoV-19/USA/NY-Wadsworth-250980-01/2020\|EPI_ISL_73 | ···················· | ···················· | ···················· | ···················· | ···················· | ···················· |
| Y | 8918 hCoV-19/USA/NY-Wadsworth-250997-01/2020\|EPI_ISL_73 | ···················· | ···················· | ···················· | ···················· | ···················· | ···················· |
| Y | 8919 hCoV-19/USA/NY-Wadsworth-251003-01/2020\|EPI_ISL_73 | ···················· | ···················· | ···················· | ···················· | ···················· | ···················· |
| Y | 8920 hCoV-19/England/MILK-C61FF5/2020\|EPI_ISL_735520\|20 | ···················· | ···················· | ···················· | ···················· | ···················· | ···················· |
| Y | 8921 hCoV-19/England/MILK-C63755/2020\|EPI_ISL_735521\|20 | ···················· | ···················· | ···················· | ···················· | ···················· | ···················· |
| Y | 8922 hCoV-19/England/MILK-C61256/2020\|EPI_ISL_735534\|20 | ···················· | ·A·················· | ···················· | ···················· | ···················· | ···A················ |
| Y | 8923 hCoV-19/England/MILK-C61362/2020\|EPI_ISL_735560\|20 | ···················· | ···················· | ···················· | ···················· | ···················· | ···················· |
| Y | 8924 hCoV-19/England/MILK-C61EE9/2020\|EPI_ISL_735563\|20 | ···················· | ···················· | ···················· | ···················· | ···················· | ···················· |
| Y | 8925 hCoV-19/England/MILK-C61A12/2020\|EPI_ISL_735565\|20 | ···················· | ···················· | ···················· | ···················· | ···················· | ···················· |
| Y | 8926 hCoV-19/England/MILK-C617EE/2020\|EPI_ISL_735567\|20 | ···················· | ···················· | ···················· | ···················· | ···················· | ···················· |
| Y | 8927 hCoV-19/England/MILK-C63904/2020\|EPI_ISL_735572\|20 | ···················· | ···················· | ···················· | ···················· | ···················· | ···················· |
| Y | 8928 hCoV-19/England/MILK-C61FD7/2020\|EPI_ISL_735574\|20 | ···················· | ···················· | ···················· | ···················· | ···················· | ···················· |
| Y | 8929 hCoV-19/England/MILK-C62D27/2020\|EPI_ISL_735576\|20 | ···················· | ·A·················· | ···················· | ···················· | ···················· | ···A················ |
| Y | 8930 hCoV-19/England/MILK-C61283/2020\|EPI_ISL_735577\|20 | ···················· | ···················· | ···················· | ···················· | ···················· | ···················· |
| Y | 8931 hCoV-19/England/MILK-C61502/2020\|EPI_ISL_735582\|20 | ···················· | ···················· | ···················· | ···················· | ···················· | ···················· |
| Y | 8932 hCoV-19/England/MILK-C6156C/2020\|EPI_ISL_735583\|20 | ···················· | ···················· | ···················· | ···················· | ···················· | ···················· |
| Y | 8933 hCoV-19/England/MILK-C62CFD/2020\|EPI_ISL_735589\|20 | ···················· | ···················· | ···················· | ···················· | ···················· | ···················· |
| Y | 8934 hCoV-19/England/MILK-C6398C/2020\|EPI_ISL_735590\|20 | ···················· | ···················· | ···················· | ···················· | ···················· | ···················· |
| Y | 8935 hCoV-19/England/MILK-C61EF8/2020\|EPI_ISL_735591\|20 | ···················· | ···················· | ···················· | ···················· | ···················· | ···················· |
| Y | 8936 hCoV-19/England/MILK-C615A8/2020\|EPI_ISL_735592\|20 | ···················· | ·A·················· | ···················· | ···················· | ···················· | ···A················ |
| Y | 8937 hCoV-19/England/MILK-C61DCE/2020\|EPI_ISL_735598\|20 | ···················· | ·A·················· | ···················· | ···················· | ···················· | ···A················ |
| Y | 8938 hCoV-19/England/MILK-C63922/2020\|EPI_ISL_735599\|20 | ···················· | ···················· | ···················· | ···················· | ······T············· | ···················· |
| Y | 8939 hCoV-19/Italy/CAM-TIGEM-108/2020\|EPI_ISL_736848\|20 | ···················· | ···A················ | ···················· | ···················· | ···················· | ···················· |
| Y | 8940 hCoV-19/Italy/CAM-TIGEM-111/2020\|EPI_ISL_736849\|20 | ···················· | ·A·················· | ···················· | ···················· | ···················· | ···················· |
| Y | 8941 hCoV-19/Italy/CAM-TIGEM-112/2020\|EPI_ISL_736850\|20 | ···················· | ···················· | ···················· | ···················· | ···················· | ···················· |
| Y | 8942 hCoV-19/Italy/CAM-TIGEM-113/2020\|EPI_ISL_736851\|20 | ···················· | ···················· | ···················· | ···················· | ···················· | ···················· |
| Y | 8943 hCoV-19/Italy/CAM-TIGEM-116/2020\|EPI_ISL_736852\|20 | ···················· | ···················· | ···················· | ···················· | ···················· | ···················· |
| Y | 8944 hCoV-19/Italy/CAM-TIGEM-118/2020\|EPI_ISL_736853\|20 | ···················· | ···················· | ···················· | ···················· | ···················· | ···················· |
| Y | 8945 hCoV-19/Italy/CAM-TIGEM-119/2020\|EPI_ISL_736854\|20 | ···················· | ···················· | ···················· | ···················· | ···················· | ···················· |
| Y | 8946 hCoV-19/Italy/CAM-TIGEM-121/2020\|EPI_ISL_736855\|20 | ···················· | ···················· | ···················· | ···················· | ···················· | ···················· |
| Y | 8947 hCoV-19/Italy/CAM-TIGEM-122/2020\|EPI_ISL_736856\|20 | ···················· | ···················· | ···················· | ···················· | ···················· | ···················· |
| Y | 8948 hCoV-19/Italy/CAM-TIGEM-123/2020\|EPI_ISL_736857\|20 | ···················· | ···················· | ···················· | ···················· | ···················· | ···················· |
| Y | 8949 hCoV-19/Italy/CAM-TIGEM-124/2020\|EPI_ISL_736858\|20 | ···················· | ···················· | ···················· | ···················· | ······T············· | ···················· |
| Y | 8950 hCoV-19/Italy/CAM-TIGEM-127/2020\|EPI_ISL_736860\|20 | ···················· | ···················· | ···················· | ···················· | ···················· | ···················· |
| Y | 8951 hCoV-19/Italy/CAM-TIGEM-130/2020\|EPI_ISL_736861\|20 | ···················· | ···················· | ···················· | ···················· | ···················· | ···················· |
| Y | 8952 hCoV-19/Italy/CAM-TIGEM-177/2020\|EPI_ISL_736890\|20 | ···················· | ···················· | ···················· | ···················· | ···················· | ···················· |
| Y | 8953 hCoV-19/Japan/IC-0420/2020\|EPI_ISL_736892\|2020-12 | ···················· | ···················· | ···················· | ···················· | ···················· | ···················· |
| Y | 8954 hCoV-19/Japan/IC-0397/2020\|EPI_ISL_736893\|2020-12 | ···················· | ·A·················· | ···················· | ···················· | ···················· | ···················· |
| Y | 8955 hCoV-19/Japan/IC-0385/2020\|EPI_ISL_736894\|2020-12 | ···················· | ···················· | ···················· | ···················· | ···················· | ···················· |
| Y | 8956 hCoV-19/Japan/IC-0401/2020\|EPI_ISL_736895\|2020-12 | ···················· | ···················· | ···················· | ···················· | ···················· | ···················· |
| Y | 8957 hCoV-19/Japan/IC-0414/2020\|EPI_ISL_736896\|2020-12 | ···················· | ···················· | ···················· | ···················· | ···················· | ···················· |
| Y | 8958 hCoV-19/Japan/IC-0386/2020\|EPI_ISL_736897\|2020-12 | ···················· | ···················· | ···················· | ···················· | ···················· | ···················· |
| Y | 8959 hCoV-19/Japan/IC-0387/2020\|EPI_ISL_736898\|2020-12 | ···················· | ···················· | ···················· | ···················· | ···················· | ···················· |
| Y | 8960 hCoV-19/Japan/IC-0388/2020\|EPI_ISL_736899\|2020-12 | ···················· | ···················· | ···················· | ···················· | ···················· | ···················· |
| Y | 8961 hCoV-19/England/NORT-1B72A49/2020\|EPI_ISL_724910\|2 | ···················· | ···················· | ···················· | ···················· | ···················· | ···················· |
| Y | 8962 hCoV-19/England/NORT-1B72B82/2020\|EPI_ISL_724915\|2 | ···················· | ···················· | ···················· | ···················· | ···················· | ···················· |
| Y | 8963 hCoV-19/England/NORT-1B72C07/2020\|EPI_ISL_724918\|2 | ···················· | ···················· | ···················· | ···················· | ···················· | ···················· |
| Y | 8964 hCoV-19/England/NORT-1B79C1F/2020\|EPI_ISL_724924\|2 | ···················· | ···················· | ···················· | ···················· | ···················· | ···················· |
| Y | 8965 hCoV-19/England/NORT-1B79C79/2020\|EPI_ISL_724925\|2 | ···················· | ···················· | ···················· | ···················· | ···················· | ···················· |
| Y | 8966 hCoV-19/England/NORT-1B79C97/2020\|EPI_ISL_724926\|2 | ···················· | ···················· | ···················· | ···················· | ···················· | ···················· |
| Y | 8967 hCoV-19/England/NORT-1B79D0D/2020\|EPI_ISL_724928\|2 | ···················· | ···················· | ···················· | ···················· | ···················· | ···················· |
| Y | 8968 hCoV-19/England/NORT-1B79D85/2020\|EPI_ISL_724932\|2 | ···················· | ···················· | ···················· | ···················· | ···················· | ···················· |
| Y | 8969 hCoV-19/England/NORT-1B79DFE/2020\|EPI_ISL_724936\|2 | ···················· | ···················· | ···················· | ···················· | ···················· | ···················· |
| Y | 8970 hCoV-19/England/NORT-1B79E37/2020\|EPI_ISL_724938\|2 | ···················· | ···················· | ···················· | ···················· | ···················· | ···················· |
| Y | 8971 hCoV-19/England/NORT-1B79E91/2020\|EPI_ISL_724939\|2 | ···················· | ···················· | ···················· | ···················· | ···················· | ···················· |
| Y | 8972 hCoV-19/England/NORT-1B79EA0/2020\|EPI_ISL_724940\|2 | ···················· | ···················· | ···················· | ···················· | ···················· | ···················· |
| Y | 8973 hCoV-19/England/NORT-1B79EEC/2020\|EPI_ISL_724941\|2 | ···················· | ···················· | ···················· | ···················· | ···················· | ···················· |
| Y | 8974 hCoV-19/England/NORT-1B79EFB/2020\|EPI_ISL_724942\|2 | ···················· | ···················· | ···················· | ···················· | ···················· | ···················· |
| Y | 8975 hCoV-19/England/NORT-1B7A003/2020\|EPI_ISL_724949\|2 | ···················· | ···················· | ···················· | ···················· | ···················· | ···················· |
| Y | 8976 hCoV-19/England/NORT-1B7A021/2020\|EPI_ISL_724950\|2 | ···················· | ···················· | ···················· | ···················· | ···················· | ···················· |
| Y | 8977 hCoV-19/England/NORT-1B7A05E/2020\|EPI_ISL_724951\|2 | ···················· | ···················· | ···················· | ···················· | ···················· | ···················· |
| Y | 8978 hCoV-19/England/NORT-1B7A0C7/2020\|EPI_ISL_724954\|2 | ···················· | ···················· | ···················· | ···················· | ···················· | ···················· |
| Y | 8979 hCoV-19/England/NORT-1B7A0F4/2020\|EPI_ISL_724955\|2 | ···················· | ···················· | ···················· | ···················· | ···················· | ···················· |
| Y | 8980 hCoV-19/England/NORT-1B7A14C/2020\|EPI_ISL_724957\|2 | ···················· | ···················· | ···················· | ···················· | ···················· | ···················· |
| Y | 8981 hCoV-19/England/NORT-1B7A16A/2020\|EPI_ISL_724958\|2 | ···················· | ···················· | ···················· | ···················· | ···················· | ···················· |
| Y | 8982 hCoV-19/England/NORT-1B7A179/2020\|EPI_ISL_724959\|2 | ···················· | ···················· | ···················· | ···················· | ···················· | ···················· |
| Y | 8983 hCoV-19/England/NORT-1B871C4/2020\|EPI_ISL_724967\|2 | ···················· | ···················· | ···················· | ···················· | ···················· | ···················· |
| Y | 8984 hCoV-19/Wales/PHWC-4986AD/2020\|EPI_ISL_727110\|2020 | ···················· | ···················· | ···················· | ···················· | ···················· | ···················· |
| Y | 8987 hCoV-19/Wales/PHWC-4986DA/2020\|EPI_ISL_727113\|2020 | ···················· | ···················· | ···················· | ···················· | ···················· | ···················· |
| Y | 8990 hCoV-19/Wales/PHWC-49875F/2020\|EPI_ISL_727118\|2020 | ···················· | ···················· | ···················· | ···················· | ···················· | ···················· |
| Y | 8991 hCoV-19/Wales/PHWC-49876E/2020\|EPI_ISL_727119\|2020 | ···················· | ···················· | ···················· | ···················· | ···················· | ···················· |

| | | | | | | | | |
|---|---|---|---|---|---|---|---|---|
| Y | 8993 hCoV-19/Wales/PHWC-4988F2/2020\|EPI_ISL_727141\|2020 | .................... | .................... | .................... | .................... | .................... | .................... | |
| Y | 8994 hCoV-19/Wales/PHWC-49890E/2020\|EPI_ISL_727142\|2020 | .................... | .................... | .................... | .................... | .................... | .................... | |
| Y | 8997 hCoV-19/Wales/PHWC-49893B/2020\|EPI_ISL_727145\|2020 | .................... | .................... | .................... | .................... | .................... | .................... | |
| Y | 9002 hCoV-19/Wales/PHWC-498995/2020\|EPI_ISL_727151\|2020 | .................... | .................... | .................... | .................... | .................... | .................... | |
| Y | 9004 hCoV-19/Wales/PHWC-4989C2/2020\|EPI_ISL_727154\|2020 | .................... | .................... | .................... | .................... | .................... | .................... | |
| Y | 9005 hCoV-19/Wales/PHWC-4989E0/2020\|EPI_ISL_727156\|2020 | .................... | .................... | .................... | .................... | .................... | .................... | |
| Y | 9006 hCoV-19/Wales/PHWC-4989FF/2020\|EPI_ISL_727157\|2020 | .................... | .................... | .................... | .................... | .................... | NNN................. | 1 |
| Y | 9008 hCoV-19/Wales/PHWC-498A1A/2020\|EPI_ISL_727159\|2020 | .................... | .................... | .................... | .................... | .................... | .................... | |
| Y | 9009 hCoV-19/Wales/PHWC-498A29/2020\|EPI_ISL_727160\|2020 | .................... | .................... | .................... | .................... | .................... | .................... | |
| Y | 9010 hCoV-19/Wales/PHWC-498A38/2020\|EPI_ISL_727161\|2020 | .................... | .................... | .................... | .................... | .................... | .................... | |
| Y | 9011 hCoV-19/Wales/PHWC-498A47/2020\|EPI_ISL_727162\|2020 | .................... | .................... | .................... | .................... | .................... | .................... | |
| Y | 9012 hCoV-19/Wales/PHWC-498A56/2020\|EPI_ISL_727163\|2020 | .................... | .................... | .................... | .................... | .................... | .................... | |
| Y | 9014 hCoV-19/Wales/PHWC-498A83/2020\|EPI_ISL_727166\|2020 | .................... | .................... | .................... | .................... | .................... | .................... | |
| Y | 9018 hCoV-19/Wales/PHWC-498B26/2020\|EPI_ISL_727176\|2020 | .................... | .................... | .................... | .................... | .................... | .................... | |
| Y | 9021 hCoV-19/Wales/PHWC-498B9F/2020\|EPI_ISL_727180\|2020 | .................... | .................... | .................... | .................... | .................... | .................... | |
| Y | 9022 hCoV-19/Wales/PHWC-498BBD/2020\|EPI_ISL_727182\|2020 | .................... | .................... | .................... | .................... | NNN................. | .................... | 1 |
| Y | 9023 hCoV-19/Wales/PHWC-498BCC/2020\|EPI_ISL_727183\|2020 | .................... | .................... | .................... | .................... | .................... | NN.......N.......... | 1 |
| Y | 9024 hCoV-19/Wales/PHWC-498BEA/2020\|EPI_ISL_727185\|2020 | .................... | .................... | .................... | .................... | NNN................. | .................... | 1 |
| Y | 9027 hCoV-19/Wales/PHWC-498C41/2020\|EPI_ISL_727191\|2020 | .................... | .................... | .................... | .................... | .................... | .................... | |
| Y | 9033 hCoV-19/USA/MI-UM-10036834038/2020\|EPI_ISL_728441\| | .................... | .................... | .................... | .................... | .................... | .................... | |
| Y | 9034 hCoV-19/USA/MI-UM-10036834498/2020\|EPI_ISL_728442\| | .................... | .................... | .................... | .................... | .................... | .................... | |
| Y | 9036 hCoV-19/USA/MI-UM-10036836150/2020\|EPI_ISL_728444\| | .................... | .................... | .................... | .................... | .................... | .................... | |
| Y | 9037 hCoV-19/USA/MI-UM-10036857948/2020\|EPI_ISL_728445\| | .................... | .................... | .................... | .................... | .................... | .................... | |
| Y | 9039 hCoV-19/USA/MI-UM-10036850527/2020\|EPI_ISL_728447\| | .................... | .................... | .................... | .................... | .................... | .................... | |
| Y | 9041 hCoV-19/USA/MI-UM-10036860017/2020\|EPI_ISL_728449\| | .................... | .................... | .................... | .................... | .................... | .................... | |
| Y | 9042 hCoV-19/USA/MI-UM-10036835591/2020\|EPI_ISL_728473\| | .................... | .................... | .................... | .................... | .................... | .................... | |
| Y | 9043 hCoV-19/USA/MI-UM-10036836394/2020\|EPI_ISL_728474\| | .................... | .................... | .................... | .................... | .................... | .................... | |
| Y | 9045 hCoV-19/USA/MI-UM-10036871873/2020\|EPI_ISL_728477\| | .................... | .................... | .................... | .................... | .................... | .................... | |
| Y | 9046 hCoV-19/USA/MI-UM-10036858347/2020\|EPI_ISL_728480\| | .................... | .................... | .................... | .................... | .................... | .................... | |
| Y | 9051 hCoV-19/USA/MI-UM-10036870465/2020\|EPI_ISL_728487\| | .................... | .................... | .................... | .................... | .................... | .................... | |
| Y | 9052 hCoV-19/USA/MI-UM-10036858179/2020\|EPI_ISL_728488\| | .................... | .................... | .................... | .................... | .................... | .................... | |
| Y | 9055 hCoV-19/USA/MI-UM-10036852009/2020\|EPI_ISL_728492\| | .................... | .................... | .................... | .................... | .................... | .................... | |
| Y | 9058 hCoV-19/USA/MI-UM-10036855736/2020\|EPI_ISL_728496\| | .................... | .................... | .................... | .................... | .................... | .................... | |
| Y | 9061 hCoV-19/England/ALDP-C3DEE6/2020\|EPI_ISL_730743\|20 | .................... | .................... | .................... | .................... | .................... | .................... | |
| Y | 9062 hCoV-19/England/ALDP-C39E9F/2020\|EPI_ISL_730747\|20 | .................... | .................... | .................... | .................... | .................... | .................... | |
| Y | 9063 hCoV-19/England/ALDP-C39D0B/2020\|EPI_ISL_730748\|20 | .................... | .................... | .................... | .................... | .................... | .................... | |
| Y | 9064 hCoV-19/England/ALDP-C3A04D/2020\|EPI_ISL_730749\|20 | .................... | .................... | .................... | .................... | .................... | .................... | |
| Y | 9065 hCoV-19/England/ALDP-C3D648/2020\|EPI_ISL_730751\|20 | .................... | .................... | .................... | .................... | .................... | .................... | |
| Y | 9066 hCoV-19/England/ALDP-C39600/2020\|EPI_ISL_730758\|20 | .................... | .................... | .................... | .................... | .................... | .................... | |
| Y | 9067 hCoV-19/England/QEUH-C30F59/2020\|EPI_ISL_730759\|20 | .................... | .................... | .................... | .................... | .................... | .................... | |
| Y | 9068 hCoV-19/England/ALDP-C3964C/2020\|EPI_ISL_730760\|20 | .................... | .................... | .................... | .................... | .................... | .................... | |
| Y | 9069 hCoV-19/England/ALDP-C39D92/2020\|EPI_ISL_730761\|20 | .................... | .................... | .................... | .................... | .................... | .................... | |
| Y | 9070 hCoV-19/England/ALDP-C3AECB/2020\|EPI_ISL_730763\|20 | .................... | .................... | .................... | .................... | .................... | .................... | |
| Y | 9071 hCoV-19/England/ALDP-C39DCF/2020\|EPI_ISL_730764\|20 | .................... | .................... | .................... | .................... | .................... | .................... | |
| Y | 9072 hCoV-19/England/ALDP-C3DE8C/2020\|EPI_ISL_730788\|20 | .................... | .................... | .................... | .................... | .................... | .................... | |
| Y | 9073 hCoV-19/England/ALDP-C39DDE/2020\|EPI_ISL_730790\|20 | .................... | .................... | .................... | .................... | .................... | .................... | |
| Y | 9074 hCoV-19/England/ALDP-C3B5F2/2020\|EPI_ISL_730791\|20 | .................... | .................... | .................... | .................... | ......T............. | .................... | |
| Y | 9075 hCoV-19/England/QEUH-C314A4/2020\|EPI_ISL_730793\|20 | .................... | .................... | .................... | .................... | .................... | .................... | |
| Y | 9076 hCoV-19/England/QEUH-C3142C/2020\|EPI_ISL_730795\|20 | .................... | .................... | .................... | .................... | .................... | .................... | |
| Y | 9077 hCoV-19/England/QEUH-C3178D/2020\|EPI_ISL_730796\|20 | .................... | .................... | .................... | .................... | .................... | .................... | |
| Y | 9078 hCoV-19/England/ALDP-C39B7A/2020\|EPI_ISL_730797\|20 | .................... | .................... | .................... | .................... | .................... | .................... | |
| Y | 9079 hCoV-19/England/ALDP-C3AC58/2020\|EPI_ISL_730798\|20 | .................... | .................... | .................... | .................... | .................... | .................... | |
| Y | 9080 hCoV-19/Russia/SVE-RII-MH239S/2020\|EPI_ISL_733002\| | .................... | .................... | .........T.......... | .................... | .................... | .................... | |
| Y | 9081 hCoV-19/Russia/SVE-RII-MH245S/2020\|EPI_ISL_733003\| | .................... | .................... | .................... | .................... | .................... | .................... | |
| Y | 9082 hCoV-19/Russia/PRI-RII-MH745S/2020\|EPI_ISL_733005\| | .................... | .................... | .................... | .................... | .................... | .................... | |
| Y | 9083 hCoV-19/Russia/PRI-RII-MH750S/2020\|EPI_ISL_733006\| | .................... | .................... | .................... | .................... | .................... | .................... | |
| Y | 9084 hCoV-19/Russia/PRI-RII-MH760S/2020\|EPI_ISL_733009\| | .................... | .................... | .........T.......... | .................... | .................... | .................... | |
| Y | 9085 hCoV-19/Russia/PRI-RII-MH762S/2020\|EPI_ISL_733010\| | .................... | .................... | .................... | .................... | .................... | .................... | |
| Y | 9086 hCoV-19/Russia/PRI-RII-MH765S/2020\|EPI_ISL_733011\| | .................... | .................... | .................... | .................... | .................... | .................... | |
| Y | 9087 hCoV-19/Russia/PRI-RII-MH766S/2020\|EPI_ISL_733012\| | .................... | .................... | .................... | .................... | .................... | .................... | |
| Y | 9088 hCoV-19/Russia/PRI-RII-MH768S/2020\|EPI_ISL_733013\| | .................... | .................... | .................... | .................... | .................... | .................... | |
| Y | 9089 hCoV-19/Russia/PRI-RII-MH829S/2020\|EPI_ISL_733018\| | ................G... | .................... | .................... | .................... | .................... | .................... | |
| Y | 9090 hCoV-19/Russia/PRI-RII-MH892S/2020\|EPI_ISL_733019\| | .................... | ....A............... | .................... | .................... | .................... | .................... | |
| Y | 9091 hCoV-19/Russia/MOW-RII-MH2884S/2020\|EPI_ISL_733053 | .................... | .................... | .................... | .................... | .................... | .................... | |
| Y | 9092 hCoV-19/Russia/MOW-RII-MH2886S/2020\|EPI_ISL_733055 | .................... | .................... | .................... | .................... | .................... | .................... | |
| Y | 9093 hCoV-19/Russia/MOW-RII-MH2896S/2020\|EPI_ISL_733059 | .................... | .................... | .................... | .................... | .................... | .................... | |
| Y | 9094 hCoV-19/USA/NY-Wadsworth-251080-01/2020\|EPI_ISL_73 | .................... | .................... | .................... | .................... | .................... | .................... | |
| Y | 9095 hCoV-19/USA/NY-Wadsworth-251095-01/2020\|EPI_ISL_73 | .................... | .................... | .................... | .................... | .................... | .................... | |
| Y | 9096 hCoV-19/USA/NY-Wadsworth-251285-01/2020\|EPI_ISL_73 | .................... | .................... | .................... | .................... | .................... | .................... | |
| Y | 9097 hCoV-19/USA/NY-Wadsworth-251999-01/2020\|EPI_ISL_73 | .................... | .................... | .................... | .................... | .................... | .................... | |
| Y | 9098 hCoV-19/USA/NY-Wadsworth-252023-01/2020\|EPI_ISL_73 | .................... | .................... | .................... | .................... | .................... | .................... | |
| Y | 9099 hCoV-19/USA/NY-Wadsworth-253098-01/2020\|EPI_ISL_73 | .................... | .................... | .................... | .................... | .................... | .................... | |
| Y | 9100 hCoV-19/USA/NY-Wadsworth-240550-01/2020\|EPI_ISL_73 | .................... | .................... | .................... | .................... | .................... | .................... | |
| Y | 9101 hCoV-19/USA/NY-Wadsworth-210267-01/2020\|EPI_ISL_73 | .................... | .................... | .................... | .................... | .................... | .................... | |
| Y | 9102 hCoV-19/USA/NY-Wadsworth-226214-01/2020\|EPI_ISL_73 | .................... | .................... | .................... | .................... | .................... | .................... | |
| Y | 9103 hCoV-19/USA/NY-Wadsworth-233280-01/2020\|EPI_ISL_73 | .................... | .A.................. | .................... | .................... | .................... | .................... | |
| Y | 9104 hCoV-19/USA/NY-Wadsworth-233575-01/2020\|EPI_ISL_73 | .................... | .............A...... | .................... | .................... | .................... | .................... | |
| Y | 9105 hCoV-19/USA/NY-Wadsworth-233695-01/2020\|EPI_ISL_73 | .................... | .............A...... | .................... | .................... | .................... | .................... | |
| Y | 9106 hCoV-19/USA/NY-Wadsworth-233888-01/2020\|EPI_ISL_73 | .................... | .................... | .................... | .................... | .................... | .................... | |
| Y | 9107 hCoV-19/USA/NY-Wadsworth-233901-01/2020\|EPI_ISL_73 | .................... | .................... | .................... | .................... | .................... | .................... | |

| | | | | | | | | |
|---|---|---|---|---|---|---|---|---|
| Y | 9108 | hCoV-19/USA/NY-Wadsworth-234464-01/2020\|EPI_ISL_73 | ···················· | ·A·················· | ····················· | ···················· | ···················· | ···················· |
| Y | 9109 | hCoV-19/USA/NY-Wadsworth-234838-01/2020\|EPI_ISL_73 | ···················· | ···················· | ····················· | ···················· | ···················· | ···················· |
| Y | 9110 | hCoV-19/USA/NY-Wadsworth-236443-01/2020\|EPI_ISL_73 | ···················· | ·A·················· | ····················· | ···················· | ···················· | ···················· |
| Y | 9111 | hCoV-19/USA/NY-Wadsworth-236998-01/2020\|EPI_ISL_73 | ···················· | ·A·················· | ····················· | ···················· | ···················· | ···················· |
| Y | 9112 | hCoV-19/USA/NY-Wadsworth-237026-01/2020\|EPI_ISL_73 | ···················· | ·A·················· | ····················· | ···················· | ···················· | ···················· |
| Y | 9113 | hCoV-19/USA/NY-Wadsworth-237064-01/2020\|EPI_ISL_73 | ···················· | ·A·················· | ····················· | ···················· | ···················· | ···················· |
| Y | 9114 | hCoV-19/USA/NY-Wadsworth-237076-01/2020\|EPI_ISL_73 | ···················· | ·A·················· | ····················· | ···················· | ···················· | ···················· |
| Y | 9115 | hCoV-19/USA/NY-Wadsworth-237082-01/2020\|EPI_ISL_73 | ···················· | ·A·················· | ····················· | ···················· | ···················· | ···················· |
| Y | 9116 | hCoV-19/USA/NY-Wadsworth-237084-01/2020\|EPI_ISL_73 | ···················· | ···················· | ····················· | ···················· | ···················· | ···················· |
| Y | 9117 | hCoV-19/USA/NY-Wadsworth-237088-01/2020\|EPI_ISL_73 | ···················· | ·A·················· | ····················· | ···················· | ···················· | ···················· |
| Y | 9118 | hCoV-19/USA/NY-Wadsworth-237092-01/2020\|EPI_ISL_73 | ···················· | ·A·················· | ····················· | ···················· | ···················· | ···················· |
| Y | 9119 | hCoV-19/USA/NY-Wadsworth-237093-01/2020\|EPI_ISL_73 | ···················· | ·A·················· | ····················· | ···················· | ···················· | ···················· |
| Y | 9120 | hCoV-19/USA/NY-Wadsworth-237102-01/2020\|EPI_ISL_73 | ···················· | ·A·················· | ····················· | ···················· | ···················· | ···················· |
| Y | 9121 | hCoV-19/USA/NY-Wadsworth-239381-01/2020\|EPI_ISL_73 | ···················· | ···················· | ····················· | ···················· | ···················· | ···················· |
| Y | 9122 | hCoV-19/USA/NY-Wadsworth-240227-01/2020\|EPI_ISL_73 | ···················· | ·A·················· | ····················· | ···················· | ···················· | ···················· |
| Y | 9123 | hCoV-19/USA/NY-Wadsworth-241177-01/2020\|EPI_ISL_73 | ···················· | ·A·················· | ····················· | ···················· | ···················· | ···················· |
| Y | 9124 | hCoV-19/USA/NY-Wadsworth-241202-01/2020\|EPI_ISL_73 | ···················· | ···················· | ····················· | ···················· | ···················· | ···················· |
| Y | 9125 | hCoV-19/USA/NY-Wadsworth-241216-01/2020\|EPI_ISL_73 | ···················· | ···················· | ····················· | ···················· | ···················· | ···················· |
| Y | 9126 | hCoV-19/USA/NY-Wadsworth-241579-01/2020\|EPI_ISL_73 | ···················· | ···················· | ····················· | ···················· | ···················· | ···················· |
| Y | 9127 | hCoV-19/USA/NY-Wadsworth-241647-01/2020\|EPI_ISL_73 | ···················· | ·A·················· | ····················· | ···················· | ···················· | ···················· |
| Y | 9128 | hCoV-19/USA/NY-Wadsworth-241671-01/2020\|EPI_ISL_73 | ···················· | ·A·················· | ····················· | ···················· | ···················· | ···················· |
| Y | 9129 | hCoV-19/USA/NY-Wadsworth-242677-01/2020\|EPI_ISL_73 | ···················· | ·A·················· | ····················· | ···················· | ···················· | ···················· |
| Y | 9130 | hCoV-19/USA/NY-Wadsworth-242678-01/2020\|EPI_ISL_73 | ···················· | ·A·················· | ····················· | ···················· | ···················· | ···················· |
| Y | 9131 | hCoV-19/USA/NY-Wadsworth-242679-01/2020\|EPI_ISL_73 | ···················· | ·A·················· | ····················· | ···················· | ···················· | ···················· |
| Y | 9132 | hCoV-19/USA/NY-Wadsworth-242689-01/2020\|EPI_ISL_73 | ···················· | ·A·················· | ····················· | ···················· | ···················· | ···················· |
| Y | 9133 | hCoV-19/USA/NY-Wadsworth-242694-01/2020\|EPI_ISL_73 | ···················· | ·A·················· | ····················· | ···················· | ···················· | ···················· |
| Y | 9134 | hCoV-19/USA/NY-Wadsworth-242695-01/2020\|EPI_ISL_73 | ···················· | ·A·················· | ····················· | ···················· | ···················· | ···················· |
| Y | 9135 | hCoV-19/USA/NY-Wadsworth-242700-01/2020\|EPI_ISL_73 | ···················· | ·A·················· | ····················· | ···················· | ···················· | ···················· |
| Y | 9136 | hCoV-19/USA/NY-Wadsworth-242707-01/2020\|EPI_ISL_73 | ···················· | ·A·················· | ····················· | ···················· | ···················· | ···················· |
| Y | 9137 | hCoV-19/USA/NY-Wadsworth-242708-01/2020\|EPI_ISL_73 | ···················· | ·A·················· | ····················· | ···················· | ···················· | ···················· |
| Y | 9138 | hCoV-19/USA/NY-Wadsworth-242716-01/2020\|EPI_ISL_73 | ···················· | ·A·················· | ····················· | ···················· | ···················· | ···················· |
| Y | 9139 | hCoV-19/USA/NY-Wadsworth-242752-01/2020\|EPI_ISL_73 | ···················· | ·A·················· | ····················· | ···················· | ···················· | ···················· |
| Y | 9140 | hCoV-19/USA/NY-Wadsworth-243231-01/2020\|EPI_ISL_73 | ···················· | ·A·················· | ····················· | ···················· | ···················· | ···················· |
| Y | 9141 | hCoV-19/USA/NY-Wadsworth-243288-01/2020\|EPI_ISL_73 | ···················· | ·A·················· | ····················· | ···················· | ···················· | ···················· |
| Y | 9142 | hCoV-19/USA/NY-Wadsworth-243296-01/2020\|EPI_ISL_73 | ···················· | ·A·················· | ····················· | ···················· | ···················· | ···················· |
| Y | 9143 | hCoV-19/USA/NY-Wadsworth-243704-01/2020\|EPI_ISL_73 | ···················· | ···················· | ····················· | ···················· | ···················· | ···················· |
| Y | 9144 | hCoV-19/USA/NY-Wadsworth-250055-01/2020\|EPI_ISL_73 | ···················· | ·A·················· | ····················· | ···················· | ···················· | ···················· |
| Y | 9145 | hCoV-19/USA/NY-Wadsworth-251287-01/2020\|EPI_ISL_73 | ···················· | ···················· | ····················· | ···················· | ···················· | ···················· |
| Y | 9146 | hCoV-19/USA/NY-Wadsworth-251302-01/2020\|EPI_ISL_73 | ···················· | ···················· | ····················· | ···················· | ···················· | ···················· |
| Y | 9147 | hCoV-19/USA/NY-Wadsworth-251307-01/2020\|EPI_ISL_73 | ···················· | ···················· | ····················· | ···················· | ···················· | ···················· |
| Y | 9148 | hCoV-19/USA/NY-Wadsworth-251308-01/2020\|EPI_ISL_73 | ···················· | ···················· | ····················· | ···················· | ···················· | ···················· |
| Y | 9149 | hCoV-19/USA/NY-Wadsworth-251314-01/2020\|EPI_ISL_73 | ···················· | ···················· | ····················· | ···················· | ···················· | ···················· |
| Y | 9150 | hCoV-19/USA/NY-Wadsworth-251316-01/2020\|EPI_ISL_73 | ···················· | ···················· | ····················· | ···················· | ···················· | ···················· |
| Y | 9151 | hCoV-19/USA/NY-Wadsworth-251320-01/2020\|EPI_ISL_73 | ···················· | ···················· | ····················· | ···················· | ···················· | ···················· |
| Y | 9152 | hCoV-19/USA/NY-Wadsworth-251942-01/2020\|EPI_ISL_73 | ···················· | ·A·················· | ····················· | ···················· | ···················· | ···················· |
| Y | 9153 | hCoV-19/USA/NY-Wadsworth-251943-01/2020\|EPI_ISL_73 | ···················· | ·A·················· | ····················· | ···················· | ···················· | ···················· |
| Y | 9154 | hCoV-19/England/MILK-C62CEE/2020\|EPI_ISL_735602\|20 | ···················· | ···················· | ···········T········· | ···················· | ···················· | ···················· |
| Y | 9155 | hCoV-19/England/MILK-C61CB2/2020\|EPI_ISL_735609\|20 | ···················· | ···················· | ····················· | ···················· | ···················· | ···················· |
| Y | 9156 | hCoV-19/England/MILK-C6249B/2020\|EPI_ISL_735615\|20 | ···················· | ···················· | ····················· | ···················· | ···················· | ···················· |
| Y | 9157 | hCoV-19/England/MILK-C61A4F/2020\|EPI_ISL_735620\|20 | ···················· | ···················· | ····················· | ···················· | ···················· | ···················· |
| Y | 9158 | hCoV-19/England/MILK-C62D09/2020\|EPI_ISL_735622\|20 | ···················· | ···················· | ····················· | ···················· | ···················· | ···················· |
| Y | 9159 | hCoV-19/England/MILK-C61B00/2020\|EPI_ISL_735626\|20 | ···················· | ···················· | ····················· | ···················· | ······T············· | ···················· |
| Y | 9160 | hCoV-19/England/MILK-C62194/2020\|EPI_ISL_735630\|20 | ···················· | ···················· | ····················· | ···················· | ···················· | ···················· |
| Y | 9161 | hCoV-19/England/MILK-C61D46/2020\|EPI_ISL_735633\|20 | ···················· | ···················· | ····················· | ···················· | ···················· | ···················· |
| Y | 9162 | hCoV-19/England/MILK-C621A3/2020\|EPI_ISL_735634\|20 | ···················· | ···················· | ····················· | ···················· | ···················· | ···················· |
| Y | 9163 | hCoV-19/England/MILK-C63A7A/2020\|EPI_ISL_735640\|20 | ···················· | ···················· | ····················· | ···················· | ···················· | ···················· |
| Y | 9164 | hCoV-19/England/MILK-C610B6/2020\|EPI_ISL_735642\|20 | ···················· | ···················· | ····················· | ···················· | ···················· | ···················· |
| Y | 9165 | hCoV-19/England/MILK-C61A21/2020\|EPI_ISL_735644\|20 | ···················· | ···················· | ····················· | ···················· | ···················· | ···················· |
| Y | 9166 | hCoV-19/England/MILK-C61ECB/2020\|EPI_ISL_735645\|20 | ···················· | ···················· | ····················· | ···················· | ···················· | ···················· |
| Y | 9167 | hCoV-19/England/MILK-C61229/2020\|EPI_ISL_735646\|20 | ···················· | ·A·················· | ····················· | ···················· | ···················· | ···A················ |
| Y | 9168 | hCoV-19/England/MILK-C6205B/2020\|EPI_ISL_735649\|20 | ···················· | ···················· | ····················· | ···················· | ······T············· | ···················· |
| Y | 9169 | hCoV-19/England/MILK-C6162D/2020\|EPI_ISL_735653\|20 | ···················· | ···················· | ····················· | ···················· | ···················· | ···················· |
| Y | 9170 | hCoV-19/England/MILK-C61E25/2020\|EPI_ISL_735656\|20 | ···················· | ···················· | ····················· | ···················· | ···················· | ···················· |
| Y | 9171 | hCoV-19/England/MILK-C61238/2020\|EPI_ISL_735657\|20 | ···················· | ·A·················· | ····················· | ···················· | ···················· | ···A················ |
| Y | 9172 | hCoV-19/England/MILK-C63263/2020\|EPI_ISL_735675\|20 | ···················· | ···················· | ····················· | ···················· | ···················· | ···················· |
| Y | 9173 | hCoV-19/England/MILK-C624E6/2020\|EPI_ISL_735676\|20 | ···················· | ···················· | ····················· | ···················· | ······T············· | ···················· |
| Y | 9174 | hCoV-19/Japan/IC-0389/2020\|EPI_ISL_736900\|2020-12 | ···················· | ···················· | ····················· | ···················· | ···················· | ···················· |
| Y | 9175 | hCoV-19/Japan/IC-0390/2020\|EPI_ISL_736901\|2020-12 | ···················· | ···················· | ····················· | ···················· | ···················· | ···················· |
| Y | 9176 | hCoV-19/Japan/IC-0391/2020\|EPI_ISL_736902\|2020-12 | ···················· | ·A·················· | ····················· | ···················· | ···················· | ···A················ |
| Y | 9177 | hCoV-19/Japan/IC-0392/2020\|EPI_ISL_736903\|2020-12 | ···················· | ·A·················· | ···········T········· | ···················· | ···················· | ···A················ |
| Y | 9178 | hCoV-19/Japan/IC-0393/2020\|EPI_ISL_736904\|2020-12 | ···················· | ···················· | ····················· | ···················· | ···················· | ···················· |
| Y | 9179 | hCoV-19/Japan/IC-0394/2020\|EPI_ISL_736905\|2020-12 | ···················· | ···················· | ····················· | ···················· | ···················· | ···················· |
| Y | 9180 | hCoV-19/Japan/IC-0395/2020\|EPI_ISL_736906\|2020-12 | ···················· | ···················· | ····················· | ···················· | ···················· | ···················· |
| Y | 9181 | hCoV-19/Japan/IC-0396/2020\|EPI_ISL_736907\|2020-12 | ···················· | ···················· | ····················· | ···················· | ···················· | ···················· |
| Y | 9182 | hCoV-19/Japan/IC-0398/2020\|EPI_ISL_736908\|2020-12 | ···················· | ···················· | ····················· | ···················· | ···················· | ···················· |
| Y | 9183 | hCoV-19/Japan/IC-0399/2020\|EPI_ISL_736909\|2020-12 | ···················· | ···················· | ····················· | ···················· | ···················· | ···················· |
| Y | 9184 | hCoV-19/Japan/IC-0400/2020\|EPI_ISL_736910\|2020-12 | ···················· | ·A·················· | ····················· | ···················· | ···················· | ···················· |
| Y | 9185 | hCoV-19/Japan/IC-0402/2020\|EPI_ISL_736911\|2020-12 | ···················· | ···················· | ····················· | ···················· | ···················· | ···················· |
| Y | 9186 | hCoV-19/Japan/IC-0403/2020\|EPI_ISL_736912\|2020-12 | ···················· | ···················· | ····················· | ···················· | ···················· | ···················· |
| Y | 9187 | hCoV-19/Japan/IC-0404/2020\|EPI_ISL_736913\|2020-12 | ···················· | ···················· | ····················· | ···················· | ···················· | ···················· |

| | | | | | | | |
|---|---|---|---|---|---|---|---|
| Y | 9188 hCoV-19/Japan/IC-0405/2020\|EPI_ISL_736914\|2020-12 | ···················· | ···················· | ···················· | ···················· | ···················· | ···················· |
| Y | 9189 hCoV-19/Japan/IC-0406/2020\|EPI_ISL_736915\|2020-12 | ···················· | ·A·················· | ···················· | ···················· | ···················· | ···A················ |
| Y | 9190 hCoV-19/Japan/IC-0407/2020\|EPI_ISL_736916\|2020-12 | ···················· | ···················· | ···················· | ···················· | ···················· | ···················· |
| Y | 9191 hCoV-19/Japan/IC-0409/2020\|EPI_ISL_736917\|2020-12 | ···················· | ···················· | ···················· | ···················· | ···················· | ···················· |
| Y | 9192 hCoV-19/Japan/IC-0410/2020\|EPI_ISL_736918\|2020-12 | ···················· | ···················· | ···················· | ···················· | ···················· | ···················· |
| Y | 9193 hCoV-19/Japan/IC-0411/2020\|EPI_ISL_736919\|2020-12 | ···················· | ···················· | ···················· | ···················· | ···················· | ···················· |
| Y | 9194 hCoV-19/Japan/IC-0412/2020\|EPI_ISL_736920\|2020-12 | ···················· | ···················· | ···················· | ···················· | ···················· | ···················· |
| Y | 9195 hCoV-19/Japan/IC-0415/2020\|EPI_ISL_736921\|2020-12 | ···················· | ·A·················· | ···················· | ···················· | ···················· | ···A················ |
| Y | 9196 hCoV-19/Japan/IC-0416/2020\|EPI_ISL_736922\|2020-12 | ···················· | ···················· | ···················· | ···················· | ···················· | ···················· |
| Y | 9197 hCoV-19/Japan/IC-0417/2020\|EPI_ISL_736923\|2020-12 | ···················· | ···················· | ···················· | ···················· | ···················· | ···················· |
| Y | 9198 hCoV-19/Japan/IC-0418/2020\|EPI_ISL_736924\|2020-12 | ···················· | ·A·················· | ···················· | ···················· | ···················· | ···················· |
| Y | 9199 hCoV-19/South Africa/KRISP-CH01110731/2020\|EPI_ISL | ···················· | ···················· | ···················· | ···················· | ···················· | ···················· |
| Y | 9200 hCoV-19/South Africa/KRISP-MDSH920916/2020\|EPI_ISL | ···················· | ···················· | ···················· | ···················· | ···················· | ···················· |
| Y | 9201 hCoV-19/South Africa/KRISP-WC-SA04352416/2020\|EPI_ | ···················· | ···················· | ···················· | ···················· | ···················· | ···················· |
| Y | 9203 hCoV-19/South Africa/KRISP-BH02956621/2020\|EPI_ISL | ···················· | ···················· | ···················· | ···················· | ···················· | ···················· |
| Y | 9204 hCoV-19/South Africa/KRISP-CH01110222/2020\|EPI_ISL | ···················· | ···················· | ···················· | ···················· | ···················· | ···················· |
| Y | 9205 hCoV-19/South Africa/KRISP-CH01110224/2020\|EPI_ISL | ···················· | ···················· | ···················· | ···················· | ···················· | ···················· |
| Y | 9206 hCoV-19/South Africa/KRISP-CH01110237/2020\|EPI_ISL | ···················· | ···················· | ···················· | ···················· | ···················· | ···················· |
| Y | 9207 hCoV-19/South Africa/KRISP-EB00584394/2020\|EPI_ISL | ···················· | ···················· | ···················· | ···················· | ···················· | ···················· |
| Y | 9208 hCoV-19/South Africa/KRISP-EB00584676/2020\|EPI_ISL | ···················· | ·A·················· | ···················· | ···················· | ···················· | ···················· |
| Y | 9209 hCoV-19/South Africa/KRISP-EH00430877/2020\|EPI_ISL | ···················· | ···················· | ···················· | ···················· | ···················· | ···················· |
| Y | 9210 hCoV-19/South Africa/KRISP-EH00431117/2020\|EPI_ISL | ···················· | ···················· | ···················· | ···················· | ···················· | ···················· |
| Y | 9211 hCoV-19/South Africa/KRISP-EJ00540655/2020\|EPI_ISL | ···················· | ···················· | ···················· | ···················· | ···················· | ···················· |
| Y | 9212 hCoV-19/South Africa/KRISP-GD00687523/2020\|EPI_ISL | ···················· | ···················· | ···················· | ···················· | ···················· | ···················· |
| Y | 9213 hCoV-19/South Africa/KRISP-GH01034869/2020\|EPI_ISL | ···················· | ···················· | ···················· | ···················· | ···················· | ···················· |
| Y | 9214 hCoV-19/South Africa/KRISP-MDSH920913/2020\|EPI_ISL | ···················· | ···················· | ···················· | ···················· | ···················· | ···················· |
| Y | 9215 hCoV-19/Czech Republic/NRL_10348/2020\|EPI_ISL_7369 | ···················· | ···················· | ···················· | ···················· | ···················· | ···················· |
| Y | 9216 hCoV-19/Czech Republic/NRL_10362/2020\|EPI_ISL_7369 | ···················· | ····G··············· | ···················· | ···················· | ···················· | ···················· |
| Y | 9217 hCoV-19/Wales/PHWC-498D20/2020\|EPI_ISL_727202\|2020 | ···················· | ···················· | ···················· | ···················· | ···················· | ···················· |
| Y | 9220 hCoV-19/Wales/PHWC-498D7B/2020\|EPI_ISL_727206\|2020 | ···················· | ···················· | ···················· | ···················· | ···················· | N··················· |
| Y | 9224 hCoV-19/Wales/PHWC-498DC6/2020\|EPI_ISL_727211\|2020 | ···················· | ···················· | ···················· | ···················· | ···················· | ···················· |
| Y | 9226 hCoV-19/Wales/PHWC-498DF3/2020\|EPI_ISL_727213\|2020 | ···················· | ···················· | ···················· | ···················· | ···················· | ···················· |
| Y | 9227 hCoV-19/Wales/PHWC-498E0F/2020\|EPI_ISL_727214\|2020 | ···················· | ···················· | ···················· | ···················· | ···················· | ···················· |
| Y | 9230 hCoV-19/Wales/PHWC-498E4B/2020\|EPI_ISL_727218\|2020 | ···················· | ···················· | ···················· | ···················· | ···················· | ···················· |
| Y | 9233 hCoV-19/USA/MI-UM-10036850953/2020\|EPI_ISL_728501\| | ···················· | ···················· | ···················· | ···················· | ···················· | ···················· |
| Y | 9241 hCoV-19/USA/MI-UM-10036861081/2020\|EPI_ISL_728511\| | ···················· | ···················· | ···················· | ···················· | ···················· | ···················· |
| Y | 9244 hCoV-19/France/ARA-107987/2020\|EPI_ISL_728516\|2020 | ···················· | ···················· | ···················· | ···················· | ···················· | ···················· |
| Y | 9245 hCoV-19/France/ARA-108293/2020\|EPI_ISL_728517\|2020 | ···················· | ···················· | ···················· | ···················· | ···················· | ···················· |
| Y | 9246 hCoV-19/France/ARA-108484/2020\|EPI_ISL_728518\|2020 | ···················· | ···················· | ···················· | ···················· | ···················· | ···················· |
| Y | 9247 hCoV-19/France/ARA-108580/2020\|EPI_ISL_728519\|2020 | ···················· | ···················· | ···················· | ···················· | ···················· | ···················· |
| Y | 9248 hCoV-19/France/ARA-108658/2020\|EPI_ISL_728520\|2020 | ···················· | ···················· | ···················· | ···················· | ···················· | ···················· |
| Y | 9249 hCoV-19/France/ARA-108709/2020\|EPI_ISL_728521\|2020 | ···················· | ···················· | ···················· | ···················· | ···················· | ···················· |
| Y | 9250 hCoV-19/France/ARA-108748/2020\|EPI_ISL_728522\|2020 | ···················· | ···················· | ···················· | ···················· | ···················· | ···················· |
| Y | 9251 hCoV-19/France/ARA-108867/2020\|EPI_ISL_728523\|2020 | ···················· | ···················· | ···················· | ···················· | ······T············· | ···············A···· |
| Y | 9252 hCoV-19/France/ARA-108907/2020\|EPI_ISL_728524\|2020 | ···················· | ···················· | ···················· | ···················· | ···················· | ···················· |
| Y | 9253 hCoV-19/France/ARA-108970/2020\|EPI_ISL_728525\|2020 | ···················· | ···················· | ···················· | ···················· | ···················· | ···················· |
| Y | 9254 hCoV-19/France/ARA-109096/2020\|EPI_ISL_728526\|2020 | ···················· | ···················· | ···················· | ···················· | ···················· | ···················· |
| Y | 9255 hCoV-19/France/ARA-109213/2020\|EPI_ISL_728527\|2020 | ···················· | ···················· | ···················· | ···················· | ···················· | ···················· |
| Y | 9256 hCoV-19/France/ARA-109325/2020\|EPI_ISL_728529\|2020 | ···················· | ···················· | ···················· | ···················· | ···················· | ···················· |
| Y | 9257 hCoV-19/France/ARA-109346/2020\|EPI_ISL_728530\|2020 | ···················· | ···················· | ···················· | ···················· | ···················· | ···················· |
| Y | 9258 hCoV-19/France/ARA-109382/2020\|EPI_ISL_728531\|2020 | ···················· | ···················· | ···················· | ···················· | ···················· | ···················· |
| Y | 9259 hCoV-19/France/ARA-109528/2020\|EPI_ISL_728532\|2020 | ···················· | ···················· | ···················· | ···················· | ···················· | ···················· |
| Y | 9260 hCoV-19/France/ARA-109965/2020\|EPI_ISL_728534\|2020 | ···················· | ···················· | ···················· | ···················· | ···················· | ···················· |
| Y | 9261 hCoV-19/France/ARA-109976/2020\|EPI_ISL_728536\|2020 | ···················· | ···················· | ···················· | ···················· | ···················· | ···················· |
| Y | 9262 hCoV-19/France/ARA-110022/2020\|EPI_ISL_728537\|2020 | ···················· | ···················· | ···················· | ···················· | ···················· | ···················· |
| Y | 9263 hCoV-19/France/ARA-110406/2020\|EPI_ISL_728538\|2020 | ···················· | ···················· | ···················· | ···················· | ···················· | ···················· |
| Y | 9264 hCoV-19/France/ARA-110538/2020\|EPI_ISL_728539\|2020 | ···················· | ···················· | ···················· | ···················· | ···················· | ···················· |
| Y | 9265 hCoV-19/France/ARA-110872/2020\|EPI_ISL_728540\|2020 | ···················· | ···················· | ···················· | ···················· | ···················· | ···················· |
| Y | 9266 hCoV-19/France/ARA-110928/2020\|EPI_ISL_728541\|2020 | ···················· | ···················· | ···················· | ···················· | ···················· | ···················· |
| Y | 9267 hCoV-19/France/ARA-110995/2020\|EPI_ISL_728542\|2020 | ···················· | ···················· | ···················· | ···················· | ···················· | ···················· |
| Y | 9268 hCoV-19/France/ARA-111014/2020\|EPI_ISL_728543\|2020 | ···················· | ···················· | ···················· | ···················· | ···················· | ···················· |
| Y | 9269 hCoV-19/France/ARA-111204/2020\|EPI_ISL_728544\|2020 | ···················· | ···················· | ···················· | ···················· | ···················· | ···················· |
| Y | 9270 hCoV-19/France/ARA-111219/2020\|EPI_ISL_728545\|2020 | ···················· | ···················· | ···················· | ···················· | ···················· | ···················· |
| Y | 9271 hCoV-19/France/ARA-111496/2020\|EPI_ISL_728546\|2020 | ···················· | ···················· | ···················· | ···················· | ···················· | ···················· |
| Y | 9272 hCoV-19/France/ARA-111501/2020\|EPI_ISL_728547\|2020 | ···················· | ···················· | ···················· | ···················· | ···················· | ···················· |
| Y | 9273 hCoV-19/France/ARA-111583/2020\|EPI_ISL_728548\|2020 | ···················· | ···················· | ···················· | ···················· | ···················· | ···················· |
| Y | 9274 hCoV-19/France/ARA-111608/2020\|EPI_ISL_728549\|2020 | ···················· | ···················· | ···················· | ···················· | ···················· | ···················· |
| Y | 9275 hCoV-19/Brazil/RS-6228/2020\|EPI_ISL_729800\|2020-05 | ···················· | ···················· | ···················· | ···················· | ···················· | ···················· |
| Y | 9276 hCoV-19/Brazil/RS-15270/2020\|EPI_ISL_729801\|2020-0 | ···················· | ···················· | ···················· | ···················· | ···················· | ···················· |
| Y | 9277 hCoV-19/Brazil/RS-15283/2020\|EPI_ISL_729802\|2020-0 | ···················· | ···················· | ···················· | ···················· | ···················· | ···················· |
| Y | 9278 hCoV-19/Brazil/RS-15286/2020\|EPI_ISL_729803\|2020-0 | ···················· | ···················· | ···················· | ···················· | ···················· | ···················· |
| Y | 9279 hCoV-19/Brazil/RS-2525/2020\|EPI_ISL_729804\|2020-03 | ···················· | ···················· | ···················· | ···················· | ···················· | ···················· |
| Y | 9280 hCoV-19/Brazil/RS-2528/2020\|EPI_ISL_729805\|2020-03 | ···················· | ···················· | ···················· | ···················· | ···················· | ···················· |
| Y | 9281 hCoV-19/Brazil/RS-2529/2020\|EPI_ISL_729806\|2020-03 | ···················· | ···················· | ···················· | ···················· | ···················· | ···················· |
| Y | 9282 hCoV-19/Brazil/RS-2533/2020\|EPI_ISL_729807\|2020-03 | ···················· | ······R············· | ···················· | ···················· | ···················· | ···················· |
| Y | 9283 hCoV-19/Brazil/RS-2539/2020\|EPI_ISL_729808\|2020-03 | ···················· | ···················· | ···················· | ···················· | ···················· | ···················· |
| Y | 9284 hCoV-19/Brazil/RS-2544/2020\|EPI_ISL_729809\|2020-03 | ···················· | ···················· | ···················· | ···················· | ···················· | ···················· |
| Y | 9285 hCoV-19/Brazil/RS-2546/2020\|EPI_ISL_729810\|2020-03 | ···················· | ···················· | ···················· | ···················· | ···················· | ···················· |
| Y | 9286 hCoV-19/Brazil/RS-2549/2020\|EPI_ISL_729811\|2020-03 | ···················· | ···················· | ···················· | ···················· | ···················· | ···················· |
| Y | 9287 hCoV-19/Brazil/RS-2550/2020\|EPI_ISL_729812\|2020-03 | ···················· | ···················· | ···················· | ···················· | ···················· | ···················· |

| | | | | | | | |
|---|---|---|---|---|---|---|---|
| Y | 9288 hCoV-19/Brazil/RS-2553/2020\|EPI_ISL_729813\|2020-03 | ···················· | ···················· | ···················· | ···················· | ···················· | ···················· |
| Y | 9289 hCoV-19/Brazil/RS-2554/2020\|EPI_ISL_729814\|2020-03 | ···················· | ···················· | ···················· | ···················· | ···················· | ···················· |
| Y | 9290 hCoV-19/Brazil/RS-2556/2020\|EPI_ISL_729815\|2020-03 | ···················· | ···················· | ···················· | ···················· | ···················· | ···················· |
| Y | 9291 hCoV-19/Brazil/RS-2564/2020\|EPI_ISL_729816\|2020-04 | ···················· | ···················· | ···················· | ···················· | ···················· | ···················· |
| Y | 9292 hCoV-19/Brazil/RS-2565/2020\|EPI_ISL_729817\|2020-04 | ···················· | ···················· | ···················· | ···················· | ···················· | ···················· |
| Y | 9293 hCoV-19/Brazil/RS-2567/2020\|EPI_ISL_729818\|2020-04 | ···················· | ···················· | ···················· | ···················· | ···················· | ···················· |
| Y | 9294 hCoV-19/Brazil/RS-6169/2020\|EPI_ISL_729819\|2020-03 | ···················· | ···················· | ···················· | ···················· | ···················· | ···················· |
| Y | 9295 hCoV-19/Brazil/RS-6177/2020\|EPI_ISL_729820\|2020-05 | ···················· | ···················· | ···················· | ···················· | ···················· | ···················· |
| Y | 9296 hCoV-19/Brazil/RS-6179/2020\|EPI_ISL_729821\|2020-05 | ···················· | ···················· | ···················· | ···················· | ···················· | ···················· |
| Y | 9297 hCoV-19/Brazil/RS-6183/2020\|EPI_ISL_729822\|2020-05 | ···················· | ···················· | ···················· | ···················· | ···················· | ···················· |
| Y | 9298 hCoV-19/Brazil/RS-6184/2020\|EPI_ISL_729823\|2020-05 | ···················· | ···················· | ···················· | ···················· | ···················· | ···················· |
| Y | 9299 hCoV-19/Brazil/RS-6188/2020\|EPI_ISL_729824\|2020-05 | ···················· | ···················· | ···················· | ···················· | ···················· | ···················· |
| Y | 9300 hCoV-19/Brazil/RS-6189/2020\|EPI_ISL_729825\|2020-05 | ···················· | ···················· | ···················· | ···················· | ···················· | ···················· |
| Y | 9301 hCoV-19/Brazil/RS-6190/2020\|EPI_ISL_729826\|2020-05 | ···················· | ···················· | ···················· | ···················· | ···················· | ···················· |
| Y | 9302 hCoV-19/Brazil/RS-6192/2020\|EPI_ISL_729827\|2020-05 | ···················· | ···················· | ···················· | ···················· | ···················· | ···················· |
| Y | 9303 hCoV-19/Brazil/RS-6196/2020\|EPI_ISL_729828\|2020-05 | ···················· | ···················· | ···················· | ···················· | ···················· | ···················· |
| Y | 9304 hCoV-19/Brazil/RS-6197/2020\|EPI_ISL_729829\|2020-05 | ···················· | ···················· | ···················· | ···················· | ···················· | ···················· |
| Y | 9305 hCoV-19/Brazil/RS-6198/2020\|EPI_ISL_729830\|2020-05 | ···················· | ···················· | ···················· | ···················· | ···················· | ···················· |
| Y | 9306 hCoV-19/Brazil/RS-6203/2020\|EPI_ISL_729831\|2020-05 | ···················· | ···················· | ···················· | ···················· | ···················· | ···················· |
| Y | 9307 hCoV-19/Brazil/RS-6205/2020\|EPI_ISL_729832\|2020-05 | ···················· | ···················· | ···················· | ···················· | ···················· | ···················· |
| Y | 9308 hCoV-19/Brazil/RS-6208/2020\|EPI_ISL_729833\|2020-05 | ···················· | ···················· | ···················· | ···················· | ···················· | ···················· |
| Y | 9309 hCoV-19/Brazil/RS-6213/2020\|EPI_ISL_729834\|2020-04 | ···················· | ···················· | ···················· | ···················· | ···················· | ···················· |
| Y | 9310 hCoV-19/Brazil/RS-6215/2020\|EPI_ISL_729835\|2020-05 | ···················· | ···················· | ···················· | ···················· | ···················· | ···················· |
| Y | 9311 hCoV-19/Brazil/RS-6218/2020\|EPI_ISL_729836\|2020-05 | ···················· | ···················· | ···················· | ···················· | ···················· | ···················· |
| Y | 9312 hCoV-19/Brazil/RS-6222/2020\|EPI_ISL_729837\|2020-05 | ···················· | ···················· | ···················· | ···················· | ···················· | ···················· |
| Y | 9313 hCoV-19/Brazil/RS-6227/2020\|EPI_ISL_729838\|2020-05 | ···················· | ···················· | ···················· | ···················· | ···················· | ···················· |
| Y | 9314 hCoV-19/Brazil/RS-6231/2020\|EPI_ISL_729839\|2020-05 | ···················· | ···················· | ···················· | ···················· | ···················· | ···················· |
| Y | 9315 hCoV-19/Brazil/RS-6232/2020\|EPI_ISL_729840\|2020-05 | ···················· | ···················· | ···················· | ···················· | ···················· | ···················· |
| Y | 9316 hCoV-19/Brazil/RS-6240/2020\|EPI_ISL_729841\|2020-06 | ···················· | ···················· | ···················· | ···················· | ···················· | ···················· |
| Y | 9317 hCoV-19/Brazil/RS-6241/2020\|EPI_ISL_729842\|2020-06 | ···················· | ···················· | ···················· | ···················· | ···················· | ···················· |
| Y | 9318 hCoV-19/Brazil/RS-6242/2020\|EPI_ISL_729843\|2020-05 | ···················· | ···················· | ···················· | ···················· | ···················· | ···················· |
| Y | 9319 hCoV-19/Brazil/RS-15273/2020\|EPI_ISL_729844\|2020-0 | ···················· | ···················· | ···················· | ···················· | ···················· | ···················· |
| Y | 9320 hCoV-19/Brazil/RS-15279/2020\|EPI_ISL_729845\|2020-0 | ···················· | ···················· | ···················· | ···················· | ···················· | ···················· |
| Y | 9321 hCoV-19/Brazil/RS-6226/2020\|EPI_ISL_729846\|2020-05 | ···················· | ···················· | ···················· | ···················· | ···················· | ···················· |
| Y | 9322 hCoV-19/Brazil/RS-6243/2020\|EPI_ISL_729847\|2020-06 | ···················· | ···················· | ···················· | ···················· | ···················· | ···················· |
| Y | 9323 hCoV-19/Brazil/RS-15278/2020\|EPI_ISL_729848\|2020-0 | ···················· | ···················· | ···················· | ···················· | ···················· | ···················· |
| Y | 9324 hCoV-19/Brazil/RS-15288/2020\|EPI_ISL_729849\|2020-0 | ···················· | ···················· | ···················· | ···················· | ···················· | ···················· |
| Y | 9325 hCoV-19/Brazil/RS-15274/2020\|EPI_ISL_729850\|2020-0 | ···················· | ···················· | ···················· | ···················· | ···················· | ···················· |
| Y | 9326 hCoV-19/Brazil/RS-15280/2020\|EPI_ISL_729851\|2020-0 | ···················· | ···················· | ···················· | ···················· | ···················· | ···················· |
| Y | 9327 hCoV-19/Brazil/RS-15281/2020\|EPI_ISL_729852\|2020-0 | ···················· | ···················· | ···················· | ···················· | ···················· | ···················· |
| Y | 9328 hCoV-19/Brazil/RS-15284/2020\|EPI_ISL_729853\|2020-0 | ···················· | ···················· | ···················· | ···················· | ···················· | ···················· |
| Y | 9329 hCoV-19/Brazil/RS-15289/2020\|EPI_ISL_729854\|2020-0 | ···················· | ···················· | ···················· | ···················· | ···················· | ···················· |
| Y | 9330 hCoV-19/Brazil/RS-15291/2020\|EPI_ISL_729855\|2020-0 | ···················· | ···················· | ···················· | ···················· | ···················· | ···················· |
| Y | 9331 hCoV-19/Brazil/RS-15275/2020\|EPI_ISL_729856\|2020-0 | ···················· | ···················· | ···················· | ···················· | ···················· | ···················· |
| Y | 9332 hCoV-19/Brazil/RS-15276/2020\|EPI_ISL_729857\|2020-0 | ···················· | ···················· | ···················· | ···················· | ···················· | ···················· |
| Y | 9333 hCoV-19/Brazil/RS-15282/2020\|EPI_ISL_729858\|2020-0 | ···················· | ···················· | ···················· | ···················· | ···················· | ···················· |
| Y | 9334 hCoV-19/Brazil/RS-15285/2020\|EPI_ISL_729859\|2020-0 | ···················· | ···················· | ···················· | ···················· | ···················· | ···················· |
| Y | 9335 hCoV-19/Brazil/RS-15287/2020\|EPI_ISL_729860\|2020-0 | ···················· | ···················· | ···················· | ···················· | ···················· | ···················· |
| Y | 9336 hCoV-19/Brazil/RS-15292/2020\|EPI_ISL_729861\|2020-0 | ···················· | ···················· | ···················· | ···················· | ···················· | ···················· |
| Y | 9337 hCoV-19/Peru/LIM-UPCH-0174/2020\|EPI_ISL_729862\|202 | ···················· | ···················· | ···················· | ···················· | ···················· | ···················· |
| Y | 9338 hCoV-19/Peru/LIM-UPCH-0178/2020\|EPI_ISL_729863\|202 | ···················· | ···················· | ···················· | ···················· | ···················· | ···················· |
| Y | 9339 hCoV-19/Peru/LIM-UPCH-0188/2020\|EPI_ISL_729864\|202 | ···················· | ···················· | ···················· | ···················· | ···················· | ···················· |
| Y | 9340 hCoV-19/Peru/LIM-UPCH-0201/2020\|EPI_ISL_729865\|202 | ···················· | ···················· | ···················· | ···················· | ···················· | ···················· |
| Y | 9341 hCoV-19/Peru/LIM-UPCH-0203/2020\|EPI_ISL_729866\|202 | ···················· | ···················· | ···················· | ···················· | ···················· | ···················· |
| Y | 9342 hCoV-19/Peru/LIM-UPCH-0208/2020\|EPI_ISL_729867\|202 | ···················· | ···················· | ···················· | ···················· | ···················· | ···················· |
| Y | 9343 hCoV-19/Peru/LIM-UPCH-0238/2020\|EPI_ISL_729868\|202 | ···················· | ···················· | ···················· | ···················· | ···················· | ···················· |
| Y | 9344 hCoV-19/Peru/AMZ-UPCH-0255/2020\|EPI_ISL_729869\|202 | ···················· | ···················· | ···················· | ···················· | ···················· | ···················· |
| Y | 9345 hCoV-19/Peru/AMZ-UPCH-0269/2020\|EPI_ISL_729870\|202 | ···················· | ····A·A············· | ···················· | ···················· | ···················· | ···················· |
| Y | 9346 hCoV-19/Peru/AMZ-UPCH-0277/2020\|EPI_ISL_729871\|202 | ···················· | ···················· | ···················· | ···················· | ···················· | ···················· |
| Y | 9347 hCoV-19/Peru/LIM-UPCH-0164/2020\|EPI_ISL_729872\|202 | ···················· | ·············A······ | ···················· | ···················· | ···················· | ···················· |
| Y | 9348 hCoV-19/Peru/LIM-UPCH-0165/2020\|EPI_ISL_729873\|202 | ···················· | ···················· | ···················· | ···················· | ···················· | ···················· |
| Y | 9349 hCoV-19/Peru/LIM-UPCH-0167/2020\|EPI_ISL_729874\|202 | ···················· | ···················· | ···················· | ···················· | ···················· | ···················· |
| Y | 9350 hCoV-19/Peru/LIM-UPCH-0168/2020\|EPI_ISL_729875\|202 | ···················· | ···················· | ···················· | ···················· | ···················· | ···················· |
| Y | 9351 hCoV-19/Peru/LIM-UPCH-0169/2020\|EPI_ISL_729876\|202 | ···················· | ···················· | ···················· | ···················· | ···················· | ···················· |
| Y | 9352 hCoV-19/Peru/LIM-UPCH-0170/2020\|EPI_ISL_729877\|202 | ···················· | ···················· | ···················· | ···················· | ···················· | ···················· |
| Y | 9353 hCoV-19/Peru/LIM-UPCH-0171/2020\|EPI_ISL_729878\|202 | ···················· | ···················· | ···················· | ···················· | ···················· | ···················· |
| Y | 9354 hCoV-19/Peru/LIM-UPCH-0173/2020\|EPI_ISL_729879\|202 | ···················· | ···················· | ···················· | ···················· | ···················· | ···················· |
| Y | 9355 hCoV-19/England/QEUH-C311BC/2020\|EPI_ISL_730802\|20 | ···················· | ···················· | ···················· | ···················· | ···················· | ···················· |
| Y | 9356 hCoV-19/England/QEUH-C319C3/2020\|EPI_ISL_730803\|20 | ···················· | ···················· | ···················· | ···················· | ···················· | ···················· |
| Y | 9357 hCoV-19/England/QEUH-C30DAA/2020\|EPI_ISL_730804\|20 | ···················· | ···················· | ···················· | ···················· | ···················· | ···················· |
| Y | 9358 hCoV-19/England/ALDP-C3ACC1/2020\|EPI_ISL_730807\|20 | ···················· | ···················· | ···················· | ···················· | ···················· | ···················· |
| Y | 9359 hCoV-19/England/QEUH-C34C12/2020\|EPI_ISL_730808\|20 | ···················· | ···················· | ···················· | ···················· | ···················· | ···················· |
| Y | 9360 hCoV-19/England/QEUH-C3226D/2020\|EPI_ISL_730809\|20 | ···················· | ···················· | ···················· | ···················· | ···················· | ···················· |
| Y | 9361 hCoV-19/England/ALDP-C39D83/2020\|EPI_ISL_730810\|20 | ···················· | ···················· | ···················· | ···················· | ···················· | ···················· |
| Y | 9362 hCoV-19/England/ALDP-C3B64A/2020\|EPI_ISL_730812\|20 | ···················· | ···················· | ···················· | ···················· | ···················· | ···················· |
| Y | 9363 hCoV-19/England/QEUH-C31A0C/2020\|EPI_ISL_730813\|20 | ···················· | ···················· | ···················· | ···················· | ···················· | ···················· |
| Y | 9364 hCoV-19/England/QEUH-C31486/2020\|EPI_ISL_730815\|20 | ···················· | ···················· | ···················· | ···················· | ···················· | ···A················ |
| Y | 9365 hCoV-19/England/ALDP-C3D60C/2020\|EPI_ISL_730818\|20 | ···················· | ···················· | ···················· | ···················· | ···················· | ···················· |
| Y | 9366 hCoV-19/England/ALDP-C3D8CA/2020\|EPI_ISL_730819\|20 | ···················· | ···················· | ···················· | ···················· | ···················· | ···················· |
| Y | 9367 hCoV-19/Wales/ALDP-C3A4BA/2020\|EPI_ISL_730822\|2020 | ···················· | ···················· | ···················· | ···················· | ···················· | ···················· |

| | | | | | | | | |
|---|---|---|---|---|---|---|---|---|
| Y | 9368 | hCoV-19/England/ALDP-C3DEAA/2020\|EPI_ISL_730823\|20 | ···················· | ···················· | ····················· | ···················· | ···················· | ···················· |
| Y | 9369 | hCoV-19/England/ALDP-C3DDBC/2020\|EPI_ISL_730825\|20 | ···················· | ···················· | ····················· | ···················· | ··A················· | ···················· |
| Y | 9370 | hCoV-19/England/ALDP-C3D89D/2020\|EPI_ISL_730826\|20 | ···················· | ···················· | ····················· | ···················· | ···················· | ···················· |
| Y | 9371 | hCoV-19/England/QEUH-C314C2/2020\|EPI_ISL_730828\|20 | ···················· | ···················· | ····················· | ···················· | ···················· | ···················· |
| Y | 9372 | hCoV-19/England/ALDP-C3D824/2020\|EPI_ISL_730829\|20 | ···················· | ···················· | ····················· | ···················· | ···················· | ···················· |
| Y | 9373 | hCoV-19/England/ALDP-C3DD43/2020\|EPI_ISL_730830\|20 | ···················· | ···················· | ····················· | ···················· | ···················· | ···················· |
| Y | 9374 | hCoV-19/England/QEUH-C321F7/2020\|EPI_ISL_730831\|20 | ···················· | ···················· | ····················· | ···················· | ···················· | ···················· |
| Y | 9375 | hCoV-19/England/ALDP-C3D587/2020\|EPI_ISL_730834\|20 | ···················· | ···················· | ····················· | ···················· | ···················· | ···················· |
| Y | 9376 | hCoV-19/England/ALDP-C3B62C/2020\|EPI_ISL_730835\|20 | ···················· | ···················· | ····················· | ···················· | ·····T·············· | ···················· |
| Y | 9377 | hCoV-19/England/ALDP-C3DD52/2020\|EPI_ISL_730839\|20 | ···················· | ···················· | ····················· | ···················· | ··A················· | ···················· |
| Y | 9378 | hCoV-19/England/ALDP-C39DA1/2020\|EPI_ISL_730843\|20 | ···················· | ···················· | ····················· | ···················· | ···················· | ···················· |
| Y | 9379 | hCoV-19/England/ALDP-C3D8E8/2020\|EPI_ISL_730845\|20 | ···················· | ···················· | ····················· | ···················· | ···················· | ···················· |
| Y | 9380 | hCoV-19/England/QEUH-C31143/2020\|EPI_ISL_730846\|20 | ···················· | ···················· | ····················· | ···················· | ·····T·············· | ···················· |
| Y | 9381 | hCoV-19/England/ALDP-C3D842/2020\|EPI_ISL_730848\|20 | ···················· | ···················· | ····················· | ···················· | ···················· | ···················· |
| Y | 9382 | hCoV-19/England/QEUH-C3224F/2020\|EPI_ISL_730849\|20 | ···················· | ···················· | ····················· | ···················· | ···················· | ···················· |
| Y | 9383 | hCoV-19/England/QEUH-C31152/2020\|EPI_ISL_730850\|20 | ···················· | ···················· | ····················· | ···················· | ···················· | ···················· |
| Y | 9384 | hCoV-19/England/ALDP-C3B61D/2020\|EPI_ISL_730851\|20 | ···················· | ···················· | ····················· | ···················· | ···················· | ···················· |
| Y | 9385 | hCoV-19/England/ALDP-C3A599/2020\|EPI_ISL_730853\|20 | ···················· | ···················· | ····················· | ···················· | ···················· | ···················· |
| Y | 9386 | hCoV-19/England/ALDP-C3ACB2/2020\|EPI_ISL_730857\|20 | ···················· | ···················· | ····················· | ···················· | ···················· | ···················· |
| Y | 9387 | hCoV-19/England/QEUH-C30D9B/2020\|EPI_ISL_730858\|20 | ···················· | ···················· | ····················· | ···················· | ···················· | ···················· |
| Y | 9388 | hCoV-19/England/ALDP-C3D666/2020\|EPI_ISL_730859\|20 | ···················· | ···················· | ····················· | ···················· | ···················· | ···················· |
| Y | 9389 | hCoV-19/England/QEUH-C32230/2020\|EPI_ISL_730860\|20 | ···················· | ···················· | ····················· | ···················· | ···················· | ···················· |
| Y | 9390 | hCoV-19/England/ALDP-C3D860/2020\|EPI_ISL_730861\|20 | ···················· | ···················· | ····················· | ···················· | ···················· | ···················· |
| Y | 9391 | hCoV-19/England/QEUH-C311F8/2020\|EPI_ISL_730862\|20 | ···················· | ···················· | ····················· | ···················· | ···················· | ···················· |
| Y | 9392 | hCoV-19/Russia/PRI-RII-MH931S/2020\|EPI_ISL_733022\| | ···················· | ···················· | ········T············ | ···················· | ···················· | ···················· |
| Y | 9393 | hCoV-19/Russia/SVE-RII-MH2833S/2020\|EPI_ISL_733024 | ···················· | ···················· | ····················· | ···················· | ···················· | ···················· |
| Y | 9394 | hCoV-19/Russia/SVE-RII-MH2834S/2020\|EPI_ISL_733025 | ···················· | ···················· | ····················· | ···················· | ···················· | ···················· |
| Y | 9395 | hCoV-19/Russia/SVE-RII-MH2840S/2020\|EPI_ISL_733027 | ···················· | ···················· | ····················· | ···················· | ···················· | ···················· |
| Y | 9396 | hCoV-19/Russia/SVE-RII-MH2841S/2020\|EPI_ISL_733028 | ···················· | ···················· | ····················· | ···················· | ···················· | ···················· |
| Y | 9397 | hCoV-19/Russia/SVE-RII-MH2843S/2020\|EPI_ISL_733030 | ···················· | ···················· | ····················· | ···················· | ···················· | ···················· |
| Y | 9398 | hCoV-19/Russia/SVE-RII-MH2845S/2020\|EPI_ISL_733032 | ···················· | ···················· | ····················· | ···················· | ···················· | ···················· |
| Y | 9399 | hCoV-19/Russia/SVE-RII-MH2847S/2020\|EPI_ISL_733033 | ···················· | ···················· | ····················· | ···················· | ···················· | ···················· |
| Y | 9400 | hCoV-19/Russia/SVE-RII-MH2850S/2020\|EPI_ISL_733034 | ···················· | ···················· | ····················· | ···················· | ···················· | ···················· |
| Y | 9401 | hCoV-19/Russia/TAM-RII-MH2855S/2020\|EPI_ISL_733037 | ···················· | ···················· | ····················· | ···················· | ···················· | ···················· |
| Y | 9402 | hCoV-19/Russia/TAM-RII-MH2857S/2020\|EPI_ISL_733039 | ···················· | ···················· | ····················· | ···················· | ···················· | ···················· |
| Y | 9403 | hCoV-19/Russia/CHE-RII-MH2861S/2020\|EPI_ISL_733041 | ···················· | ···················· | ····················· | ···················· | ···················· | ···················· |
| Y | 9404 | hCoV-19/Russia/MOW-RII-MH2868S/2020\|EPI_ISL_733044 | ···················· | ···················· | ····················· | ···················· | ···················· | ···················· |
| Y | 9405 | hCoV-19/Russia/MOW-RII-MH2899S/2020\|EPI_ISL_733060 | ···················· | ···················· | ····················· | ···················· | ···················· | ···················· |
| Y | 9406 | hCoV-19/Russia/MOW-RII-MH2900S/2020\|EPI_ISL_733061 | ···················· | ···················· | ····················· | ···················· | ···················· | ···················· |
| Y | 9407 | hCoV-19/Russia/MOW-RII-MH2903S/2020\|EPI_ISL_733062 | ···················· | ···················· | ····················· | ···················· | ···················· | ···················· |
| Y | 9408 | hCoV-19/Russia/MOW-RII-MH2906S/2020\|EPI_ISL_733065 | ···················· | ···················· | ····················· | ···················· | ···················· | ···················· |
| Y | 9409 | hCoV-19/Russia/MOW-RII-MH2907S/2020\|EPI_ISL_733066 | ···················· | ···················· | ····················· | ···················· | ···················· | ···················· |
| Y | 9410 | hCoV-19/Russia/MOW-RII-MH2911S/2020\|EPI_ISL_733067 | ···················· | ···················· | ····················· | ···················· | ···················· | ···················· |
| Y | 9411 | hCoV-19/Russia/MOW-RII-MH2917S/2020\|EPI_ISL_733069 | ···················· | ···················· | ····················· | ···················· | ···················· | ···················· |
| Y | 9412 | hCoV-19/Russia/MOW-RII-MH2923S/2020\|EPI_ISL_733071 | ···················· | ···················· | ····················· | ···················· | ···················· | ···················· |
| Y | 9413 | hCoV-19/Russia/MOW-RII-MH2925S/2020\|EPI_ISL_733073 | ···················· | ···················· | ····················· | ···················· | ···················· | ···················· |
| Y | 9414 | hCoV-19/Russia/MOW-RII-MH2929S/2020\|EPI_ISL_733074 | ···················· | ···················· | ····················· | ···················· | ···················· | ···················· |
| Y | 9415 | hCoV-19/Russia/KHM-RII-MH2930S/2020\|EPI_ISL_733075 | ···················· | ···················· | ····················· | ···················· | ···················· | ···················· |
| Y | 9416 | hCoV-19/Crimea/RII-MH2936S/2020\|EPI_ISL_733077\|202 | ···················· | ···················· | ····················· | ···················· | ···················· | ···················· |
| Y | 9417 | hCoV-19/Russia/NGR-RII-MH2939S/2020\|EPI_ISL_733079 | ···················· | ···················· | ····················· | ···················· | ···················· | ···················· |
| Y | 9418 | hCoV-19/Russia/SVE-RII-MH2956S/2020\|EPI_ISL_733080 | ···················· | ···················· | ····················· | ···················· | ···················· | ···················· |
| Y | 9419 | hCoV-19/Russia/SVE-RII-MH2962S/2020\|EPI_ISL_733082 | ···················· | ···················· | ····················· | ···················· | ···················· | ···················· |
| Y | 9420 | hCoV-19/Russia/SVE-RII-MH2973S/2020\|EPI_ISL_733086 | ···················· | ···················· | ····················· | ···················· | ···················· | ···················· |
| Y | 9421 | hCoV-19/Russia/SVE-RII-MH2978S/2020\|EPI_ISL_733087 | ···················· | ···················· | ····················· | ···················· | ···················· | ···················· |
| Y | 9422 | hCoV-19/Russia/SVE-RII-MH3109S/2020\|EPI_ISL_733088 | ···················· | ···················· | ····················· | ···················· | ···················· | ···················· |
| Y | 9424 | hCoV-19/Russia/SVE-RII-MH3111S/2020\|EPI_ISL_733090 | ···················· | ···················· | ····················· | ···················· | ···················· | ···················· |
| Y | 9425 | hCoV-19/Russia/TYU-RII-MH3116S/2020\|EPI_ISL_733092 | ···················· | ···················· | ····················· | ···················· | ···················· | ···················· |
| Y | 9426 | hCoV-19/Russia/LEN-RII-MH3118S/2020\|EPI_ISL_733093 | ···················· | ···················· | ····················· | ···················· | ···················· | ···················· |
| Y | 9427 | hCoV-19/Russia/LEN-RII-MH3120S/2020\|EPI_ISL_733095 | ···················· | ···················· | ····················· | ···················· | ···················· | ···················· |
| Y | 9428 | hCoV-19/Russia/LEN-RII-MH3121S/2020\|EPI_ISL_733096 | ···················· | ···················· | ····················· | ···················· | ···················· | ···················· |
| Y | 9429 | hCoV-19/Russia/LEN-RII-MH3122S/2020\|EPI_ISL_733097 | ···················· | ···················· | ····················· | ···················· | ···T·T·············· | ···················· |
| Y | 9430 | hCoV-19/USA/NY-Wadsworth-243895-01/2020\|EPI_ISL_73 | ···················· | ·A·················· | ····················· | ···················· | ···················· | ···················· |
| Y | 9431 | hCoV-19/USA/NY-Wadsworth-245457-01/2020\|EPI_ISL_73 | ···················· | ·A·················· | ····················· | ···················· | ···················· | ···················· |
| Y | 9432 | hCoV-19/USA/NY-Wadsworth-245811-01/2020\|EPI_ISL_73 | ···················· | ·A·················· | ····················· | ···················· | ···················· | ···················· |
| Y | 9433 | hCoV-19/USA/NY-Wadsworth-247244-01/2020\|EPI_ISL_73 | ···················· | ···················· | ····················· | ···················· | ···················· | ···················· |
| Y | 9434 | hCoV-19/USA/NY-Wadsworth-247373-01/2020\|EPI_ISL_73 | ···················· | ···················· | ····················· | ···················· | ···················· | ···················· |
| Y | 9435 | hCoV-19/USA/NY-Wadsworth-247518-01/2020\|EPI_ISL_73 | ···················· | ···················· | ····················· | ···················· | ···················· | ···················· |
| Y | 9436 | hCoV-19/USA/NY-Wadsworth-247525-01/2020\|EPI_ISL_73 | ···················· | ···················· | ····················· | ···················· | ···················· | ···················· |
| Y | 9437 | hCoV-19/USA/NY-Wadsworth-247528-01/2020\|EPI_ISL_73 | ···················· | ···················· | ····················· | ···················· | ···················· | ···················· |
| Y | 9438 | hCoV-19/USA/NY-Wadsworth-247569-01/2020\|EPI_ISL_73 | ···················· | ···················· | ····················· | ···················· | ···················· | ···················· |
| Y | 9439 | hCoV-19/USA/NY-Wadsworth-247571-01/2020\|EPI_ISL_73 | ···················· | ···················· | ····················· | ···················· | ···················· | ···················· |
| Y | 9440 | hCoV-19/USA/NY-Wadsworth-247575-01/2020\|EPI_ISL_73 | ···················· | ···················· | ····················· | ···················· | ···················· | ···················· |
| Y | 9441 | hCoV-19/USA/NY-Wadsworth-247826-01/2020\|EPI_ISL_73 | ···················· | ···················· | ····················· | ···················· | ···················· | ···················· |
| Y | 9442 | hCoV-19/USA/NY-Wadsworth-247837-01/2020\|EPI_ISL_73 | ···················· | ···················· | ····················· | ···················· | ···················· | ···················· |
| Y | 9443 | hCoV-19/USA/NY-Wadsworth-247867-01/2020\|EPI_ISL_73 | ···················· | ···················· | ····················· | ···················· | ···················· | ···················· |
| Y | 9444 | hCoV-19/USA/NY-Wadsworth-248007-01/2020\|EPI_ISL_73 | ···················· | ···················· | ····················· | ···················· | ···················· | ···················· |
| Y | 9445 | hCoV-19/USA/NY-Wadsworth-248011-01/2020\|EPI_ISL_73 | ···················· | ···················· | ····················· | ···················· | ···················· | ···················· |
| Y | 9446 | hCoV-19/USA/NY-Wadsworth-248016-01/2020\|EPI_ISL_73 | ···················· | ···················· | ····················· | ···················· | ···················· | ···················· |
| Y | 9447 | hCoV-19/USA/NY-Wadsworth-248102-01/2020\|EPI_ISL_73 | ···················· | ···················· | ····················· | ···················· | ···················· | ···················· |
| Y | 9448 | hCoV-19/USA/NY-Wadsworth-248513-01/2020\|EPI_ISL_73 | ···················· | ·A·················· | ····················· | ···················· | ···················· | ···················· |

| | | | | | | | |
|---|---|---|---|---|---|---|---|
| Y | 9449 hCoV-19/USA/NY-Wadsworth-248541-01/2020\|EPI_ISL_73 | ···················· | ·A·················· | ····················· | ···················· | ···················· | ···················· |
| Y | 9450 hCoV-19/USA/NY-Wadsworth-248542-01/2020\|EPI_ISL_73 | ···················· | ·A·················· | ····················· | ···················· | ···················· | ···················· |
| Y | 9451 hCoV-19/USA/NY-Wadsworth-248543-01/2020\|EPI_ISL_73 | ···················· | ·A·················· | ····················· | ···················· | ···················· | ···················· |
| Y | 9452 hCoV-19/USA/NY-Wadsworth-248647-01/2020\|EPI_ISL_73 | ···················· | ···················· | ····················· | ···················· | ···················· | ···················· |
| Y | 9453 hCoV-19/USA/NY-Wadsworth-248731-01/2020\|EPI_ISL_73 | ···················· | ···················· | ····················· | ···················· | ···················· | ···················· |
| Y | 9454 hCoV-19/USA/NY-Wadsworth-248789-01/2020\|EPI_ISL_73 | ···················· | ···················· | ····················· | ···················· | ···················· | ···················· |
| Y | 9455 hCoV-19/USA/NY-Wadsworth-248792-01/2020\|EPI_ISL_73 | ···················· | ···················· | ····················· | ···················· | ···················· | ···················· |
| Y | 9456 hCoV-19/USA/NY-Wadsworth-249752-01/2020\|EPI_ISL_73 | ···················· | ···················· | ····················· | ···················· | ···················· | ···················· |
| Y | 9457 hCoV-19/USA/NY-Wadsworth-249841-01/2020\|EPI_ISL_73 | ···················· | ···················· | ····················· | ···················· | ···················· | ···················· |
| Y | 9458 hCoV-19/USA/NY-Wadsworth-249924-01/2020\|EPI_ISL_73 | ···················· | ···················· | ····················· | ···················· | ···················· | ···················· |
| Y | 9459 hCoV-19/USA/NY-Wadsworth-249952-01/2020\|EPI_ISL_73 | ···················· | ···················· | ····················· | ···················· | ···················· | ···················· |
| Y | 9460 hCoV-19/USA/NY-Wadsworth-251944-01/2020\|EPI_ISL_73 | ···················· | ·A·················· | ····················· | ···················· | ···················· | ···················· |
| Y | 9461 hCoV-19/USA/NY-Wadsworth-251946-01/2020\|EPI_ISL_73 | ···················· | ·A·················· | ····················· | ···················· | ···················· | ···················· |
| Y | 9462 hCoV-19/USA/NY-Wadsworth-251951-01/2020\|EPI_ISL_73 | ···················· | ·A·················· | ····················· | ···················· | ···················· | ···················· |
| Y | 9463 hCoV-19/USA/NY-Wadsworth-251954-01/2020\|EPI_ISL_73 | ···················· | ·A·················· | ····················· | ···················· | ···················· | ···················· |
| Y | 9464 hCoV-19/USA/NY-Wadsworth-251958-01/2020\|EPI_ISL_73 | ···················· | ·A·················· | ····················· | ···················· | ···················· | ···················· |
| Y | 9465 hCoV-19/USA/NY-Wadsworth-251963-01/2020\|EPI_ISL_73 | ···················· | ·A·················· | ····················· | ···················· | ···················· | ···················· |
| Y | 9466 hCoV-19/USA/NY-Wadsworth-251969-01/2020\|EPI_ISL_73 | ···················· | ·A·················· | ····················· | ···················· | ···················· | ···················· |
| Y | 9467 hCoV-19/USA/NY-Wadsworth-251975-01/2020\|EPI_ISL_73 | ···················· | ·A·················· | ····················· | ···················· | ···················· | ···················· |
| Y | 9468 hCoV-19/USA/NY-Wadsworth-252108-01/2020\|EPI_ISL_73 | ···················· | ···················· | ····················· | ···················· | ···················· | ···················· |
| Y | 9469 hCoV-19/USA/NY-Wadsworth-252153-01/2020\|EPI_ISL_73 | ···················· | ···················· | ····················· | ···················· | ···················· | ···················· |
| Y | 9470 hCoV-19/England/MILK-C61E70/2020\|EPI_ISL_735688\|20 | ···················· | ···················· | ····················· | ···················· | ···················· | ···················· |
| Y | 9471 hCoV-19/England/MILK-C61FE6/2020\|EPI_ISL_735693\|20 | ···················· | ···················· | ····················· | ···················· | ···················· | ···················· |
| Y | 9472 hCoV-19/England/MILK-C6203D/2020\|EPI_ISL_735696\|20 | ···················· | ···················· | ····················· | ···················· | ···················· | ···················· |
| Z | 1 hCoV-19/England/SHEF-10CDDD8/2020\|EPI_ISL_742005\|2 | ···················· | ···················· | ····················· | ···················· | ······T············· | ···················· |
| Z | 2 hCoV-19/England/SHEF-10CFD4F/2020\|EPI_ISL_742022\|2 | ···················· | ···················· | ····················· | ···················· | ···················· | ···················· |
| Z | 3 hCoV-19/England/SHEF-10CFEF1/2020\|EPI_ISL_742024\|2 | ···················· | ···················· | ····················· | ···················· | ······T············· | ···················· |
| Z | 4 hCoV-19/England/SHEF-10D0B14/2020\|EPI_ISL_742028\|2 | ···················· | ···················· | ····················· | ···················· | ···················· | ···················· |
| Z | 5 hCoV-19/England/SHEF-10D107E/2020\|EPI_ISL_742031\|2 | ···················· | ···················· | ····················· | ···················· | ···················· | ···················· |
| Z | 6 hCoV-19/England/SHEF-10D2259/2020\|EPI_ISL_742044\|2 | ···················· | ···················· | ····················· | ···················· | ···················· | ···················· |
| Z | 7 hCoV-19/England/SHEF-10D2C97/2020\|EPI_ISL_742048\|2 | ···················· | ···················· | ····················· | ···················· | ···················· | ···················· |
| Z | 8 hCoV-19/Wales/PHWC-49F9BC/2020\|EPI_ISL_743300\|2020 | ···················· | ···················· | ····················· | ···················· | ···················· | ···················· |
| Z | 9 hCoV-19/Wales/PHWC-49F9CB/2020\|EPI_ISL_743301\|2020 | ···················· | ···················· | ····················· | ···················· | ···················· | ···················· |
| Z | 10 hCoV-19/Wales/PHWC-49F9DA/2020\|EPI_ISL_743302\|2020 | ···················· | ···················· | ····················· | ···················· | ···················· | ···················· |
| Z | 11 hCoV-19/Wales/PHWC-49F9E9/2020\|EPI_ISL_743303\|2020 | ···················· | ···················· | ····················· | ···················· | ···················· | ···················· |
| Z | 12 hCoV-19/Wales/PHWC-49F9F8/2020\|EPI_ISL_743304\|2020 | ···················· | ···················· | ····················· | ···················· | ···················· | ···················· |
| Z | 13 hCoV-19/Wales/PHWC-49FA13/2020\|EPI_ISL_743306\|2020 | ···················· | ···················· | ····················· | ···················· | ···················· | ···················· |
| Z | 14 hCoV-19/Wales/PHWC-49FA22/2020\|EPI_ISL_743307\|2020 | ···················· | ···················· | ····················· | ···················· | ···················· | ···················· |
| Z | 15 hCoV-19/Wales/PHWC-49FA31/2020\|EPI_ISL_743308\|2020 | ···················· | ···················· | ····················· | ···················· | ···················· | ···················· |
| Z | 16 hCoV-19/Wales/PHWC-49FA40/2020\|EPI_ISL_743309\|2020 | ···················· | ···················· | ····················· | ···················· | ···················· | ···················· |
| Z | 17 hCoV-19/Wales/PHWC-49FA6E/2020\|EPI_ISL_743311\|2020 | ···················· | ···················· | ····················· | ···················· | ···················· | ···················· |
| Z | 18 hCoV-19/Wales/PHWC-49FA7D/2020\|EPI_ISL_743312\|2020 | ···················· | ···················· | ····················· | ···················· | ···················· | ···················· |
| Z | 19 hCoV-19/Wales/PHWC-49FA8C/2020\|EPI_ISL_743313\|2020 | ···················· | ···················· | ····················· | ···················· | ···················· | ···················· |
| Z | 20 hCoV-19/Wales/PHWC-49FA9B/2020\|EPI_ISL_743314\|2020 | ···················· | ···················· | ····················· | ···················· | ···················· | ···················· |
| Z | 22 hCoV-19/Wales/PHWC-49FAB9/2020\|EPI_ISL_743316\|2020 | ···················· | ···················· | ····················· | ···················· | ···················· | ···················· |
| Z | 23 hCoV-19/Wales/PHWC-49FAC8/2020\|EPI_ISL_743317\|2020 | ···················· | ···················· | ····················· | ···················· | ···················· | ···················· |
| Z | 24 hCoV-19/Wales/PHWC-49FAD7/2020\|EPI_ISL_743318\|2020 | ···················· | ···················· | ····················· | ···················· | ···················· | ···················· |
| Z | 25 hCoV-19/Wales/PHWC-49FAE6/2020\|EPI_ISL_743319\|2020 | ···················· | ···················· | ····················· | ···················· | ···················· | ···················· |
| Z | 26 hCoV-19/Wales/PHWC-49FAF5/2020\|EPI_ISL_743320\|2020 | ···················· | ···················· | ····················· | ···················· | ···················· | ···················· |
| Z | 27 hCoV-19/Wales/PHWC-49FB01/2020\|EPI_ISL_743321\|2020 | ···················· | ···················· | ····················· | ···················· | ···················· | ···················· |
| Z | 28 hCoV-19/Wales/PHWC-49FB3E/2020\|EPI_ISL_743322\|2020 | ···················· | ···················· | ····················· | ···················· | ···················· | ···················· |
| Z | 29 hCoV-19/Wales/PHWC-49FB6B/2020\|EPI_ISL_743324\|2020 | ···················· | ···················· | ····················· | ···················· | ···················· | ···················· |
| Z | 30 hCoV-19/Wales/PHWC-49FB7A/2020\|EPI_ISL_743325\|2020 | ···················· | ···················· | ····················· | ···················· | ···················· | ···················· |
| Z | 31 hCoV-19/Wales/PHWC-49FBA7/2020\|EPI_ISL_743327\|2020 | ···················· | ···················· | ····················· | ···················· | ···················· | ···················· |
| Z | 33 hCoV-19/Wales/PHWC-49FBC5/2020\|EPI_ISL_743329\|2020 | ···················· | ···················· | ····················· | ···················· | ···················· | ···················· |
| Z | 34 hCoV-19/Wales/PHWC-49FBD4/2020\|EPI_ISL_743330\|2020 | ···················· | ···················· | ····················· | ···················· | ···················· | ···················· |
| Z | 35 hCoV-19/Wales/PHWC-49FBE3/2020\|EPI_ISL_743331\|2020 | ···················· | ···················· | ····················· | ···················· | ···················· | ···················· |
| Z | 36 hCoV-19/Wales/PHWC-49FC0E/2020\|EPI_ISL_743333\|2020 | ···················· | ···················· | ····················· | ···················· | ···················· | ···················· |
| Z | 37 hCoV-19/Wales/PHWC-49FC1D/2020\|EPI_ISL_743334\|2020 | ···················· | ···················· | ····················· | ···················· | ···················· | ···················· |
| Z | 38 hCoV-19/Wales/PHWC-49FC2C/2020\|EPI_ISL_743335\|2020 | ···················· | ···················· | ····················· | ···················· | ···················· | ···················· |
| Z | 39 hCoV-19/Wales/PHWC-49FC4A/2020\|EPI_ISL_743336\|2020 | ···················· | ···················· | ····················· | ···················· | ···················· | ···················· |
| Z | 40 hCoV-19/Wales/PHWC-49FC59/2020\|EPI_ISL_743337\|2020 | ···················· | ···················· | ····················· | ···················· | ···················· | ···················· |
| Z | 41 hCoV-19/Wales/PHWC-49FC68/2020\|EPI_ISL_743338\|2020 | ···················· | ···················· | ····················· | ···················· | ···················· | ···················· |
| Z | 42 hCoV-19/Wales/PHWC-49FCA4/2020\|EPI_ISL_743340\|2020 | ···················· | ···················· | ····················· | ···················· | ···················· | ···················· |
| Z | 43 hCoV-19/Wales/PHWC-49FCB3/2020\|EPI_ISL_743341\|2020 | ···················· | ···················· | ····················· | ···················· | ···················· | ···················· |
| Z | 44 hCoV-19/Wales/PHWC-49FCC2/2020\|EPI_ISL_743342\|2020 | ···················· | ···················· | ····················· | ···················· | ···················· | ···················· |
| Z | 46 hCoV-19/Wales/PHWC-49FCFF/2020\|EPI_ISL_743345\|2020 | ···················· | ···················· | ····················· | ···················· | ···················· | ···················· |
| Z | 47 hCoV-19/Wales/PHWC-49FD0B/2020\|EPI_ISL_743346\|2020 | ···················· | ···················· | ····················· | ···················· | ···················· | ···················· |
| Z | 49 hCoV-19/Wales/PHWC-49FD38/2020\|EPI_ISL_743348\|2020 | ···················· | ···················· | ····················· | ···················· | ···················· | ···················· |
| Z | 51 hCoV-19/Wales/PHWC-49FD56/2020\|EPI_ISL_743350\|2020 | ···················· | ···················· | ····················· | ···················· | ···················· | ···················· |
| Z | 53 hCoV-19/Wales/PHWC-49FDED/2020\|EPI_ISL_743357\|2020 | ···················· | ···················· | ····················· | ···················· | ···················· | ···················· |
| Z | 56 hCoV-19/Wales/PHWC-49FECC/2020\|EPI_ISL_743365\|2020 | ···················· | ···················· | ····················· | ···················· | ···················· | ···················· |
| Z | 57 hCoV-19/Wales/PHWC-49FEDB/2020\|EPI_ISL_743366\|2020 | ···················· | ···················· | ····················· | ···················· | ···················· | ···················· |
| Z | 58 hCoV-19/Wales/PHWC-49FEF9/2020\|EPI_ISL_743368\|2020 | ···················· | ···················· | ····················· | ···················· | ···················· | ···················· |
| Z | 62 hCoV-19/Wales/PHWC-49FF41/2020\|EPI_ISL_743372\|2020 | ···················· | ···················· | ····················· | ···················· | ···················· | ···················· |
| Z | 63 hCoV-19/Wales/PHWC-49FF50/2020\|EPI_ISL_743373\|2020 | ···················· | ···················· | ····················· | ···················· | ···················· | ···················· |
| Z | 64 hCoV-19/Wales/PHWC-49FF7E/2020\|EPI_ISL_743375\|2020 | ···················· | ···················· | ····················· | ···················· | ···················· | ···················· |
| Z | 65 hCoV-19/Wales/PHWC-49FF8D/2020\|EPI_ISL_743376\|2020 | ···················· | ···················· | ····················· | ···················· | ···················· | ···················· |
| Z | 66 hCoV-19/Wales/PHWC-49FFAB/2020\|EPI_ISL_743378\|2020 | ···················· | ···················· | ····················· | ···················· | ···················· | ···················· |
| Z | 67 hCoV-19/Wales/PHWC-49FFBA/2020\|EPI_ISL_743379\|2020 | ···················· | ···················· | ····················· | ···················· | ···················· | ···················· |

| | | | | | | | | |
|---|---|---|---|---|---|---|---|---|
| Z | 68 | hCoV-19/Luxembourg/LNS7627646/2020\|EPI_ISL_744600\| | ···················· | ···················· | ···················· | ···················· | ···················· | ···················· |
| Z | 69 | hCoV-19/Luxembourg/LNS7628344/2020\|EPI_ISL_744601\| | ···················· | ···················· | ···················· | ···················· | ·····T·············· | ···················· |
| Z | 70 | hCoV-19/Luxembourg/LNS7630398/2020\|EPI_ISL_744602\| | ···················· | ···················· | ···················· | ···················· | ·····T·············· | ···················· |
| Z | 71 | hCoV-19/Luxembourg/LNS7632775/2020\|EPI_ISL_744603\| | ···················· | ···················· | ···················· | ···················· | ···················· | ···················· |
| Z | 72 | hCoV-19/Luxembourg/LNS7638444/2020\|EPI_ISL_744604\| | ···················· | ···················· | ···················· | ···················· | ·····T·············· | ···················· |
| Z | 73 | hCoV-19/Luxembourg/LNS7642204/2020\|EPI_ISL_744605\| | ···················· | ···················· | ···················· | ···················· | ···················· | ···················· |
| Z | 74 | hCoV-19/Luxembourg/LNS7643222/2020\|EPI_ISL_744606\| | ···················· | ···················· | ···················· | ···················· | ···················· | ···················· |
| Z | 75 | hCoV-19/Luxembourg/LNS7658808/2020\|EPI_ISL_744608\| | ···················· | ···················· | ···················· | ···················· | ···················· | ···················· |
| Z | 76 | hCoV-19/Luxembourg/LNS7680274/2020\|EPI_ISL_744611\| | ···················· | ···················· | ···················· | ···················· | ···················· | ···················· |
| Z | 77 | hCoV-19/Luxembourg/LNS7680363/2020\|EPI_ISL_744612\| | ···················· | ·A·················· | ···················· | ···················· | ···················· | ···················· |
| Z | 78 | hCoV-19/Luxembourg/LNS7680586/2020\|EPI_ISL_744613\| | ···················· | ···················· | ···················· | ···················· | ···················· | ···················· |
| Z | 79 | hCoV-19/Luxembourg/LNS7686105/2020\|EPI_ISL_744614\| | ···················· | ···················· | ···················· | ···················· | ···················· | ···················· |
| Z | 80 | hCoV-19/Luxembourg/LNS7686862/2020\|EPI_ISL_744615\| | ···················· | ···················· | ···················· | ···················· | ···················· | ···················· |
| Z | 81 | hCoV-19/Luxembourg/LNS7693582/2020\|EPI_ISL_744616\| | ···················· | ···················· | ···················· | ···················· | ···················· | ···················· |
| Z | 82 | hCoV-19/Luxembourg/LNS7693672/2020\|EPI_ISL_744617\| | ···················· | ···················· | ···················· | ···················· | ···················· | ···················· |
| Z | 83 | hCoV-19/Luxembourg/LNS7716837/2020\|EPI_ISL_744619\| | ···················· | ···················· | ···················· | ···················· | ···················· | ···················· |
| Z | 84 | hCoV-19/Luxembourg/LNS7719161/2020\|EPI_ISL_744620\| | ···················· | ···················· | ···················· | ···················· | ···················· | ···················· |
| Z | 85 | hCoV-19/Luxembourg/LNS7724491/2020\|EPI_ISL_744621\| | ···················· | ···················· | ···················· | ···················· | ···················· | ···················· |
| Z | 86 | hCoV-19/Luxembourg/LNS7727746/2020\|EPI_ISL_744622\| | ···················· | ···················· | ···················· | ···················· | ···················· | ···················· |
| Z | 87 | hCoV-19/Luxembourg/LNS7729675/2020\|EPI_ISL_744623\| | ···················· | ···················· | ···················· | ···················· | ···················· | ···················· |
| Z | 88 | hCoV-19/Luxembourg/LNS7741644/2020\|EPI_ISL_744624\| | ···················· | ···················· | ···················· | ···················· | ···················· | ···················· |
| Z | 89 | hCoV-19/Luxembourg/LNS7744413/2020\|EPI_ISL_744625\| | ···················· | ···················· | ···················· | ···················· | ···················· | ···················· |
| Z | 90 | hCoV-19/Luxembourg/LNS7745026/2020\|EPI_ISL_744626\| | ···················· | ···················· | ···················· | ···················· | ···················· | ···················· |
| Z | 91 | hCoV-19/Luxembourg/LNS7748234/2020\|EPI_ISL_744627\| | ···················· | ···················· | ···················· | ···················· | ···················· | ···················· |
| Z | 92 | hCoV-19/Luxembourg/LNS7760954/2020\|EPI_ISL_744628\| | ···················· | ···················· | ···················· | ···················· | ···················· | ···················· |
| Z | 93 | hCoV-19/Luxembourg/LNS7765858/2020\|EPI_ISL_744629\| | ···················· | ···················· | ···················· | ···················· | ···················· | ···················· |
| Z | 94 | hCoV-19/Luxembourg/LNS7775195/2020\|EPI_ISL_744630\| | ···················· | ···················· | ···················· | ···················· | ···················· | ···················· |
| Z | 95 | hCoV-19/Luxembourg/LNS7788775/2020\|EPI_ISL_744631\| | ···················· | ···················· | ···················· | ···················· | ···················· | ···················· |
| Z | 96 | hCoV-19/Luxembourg/LNS7793491/2020\|EPI_ISL_744632\| | ···················· | ···················· | ···················· | ···················· | ···················· | ···················· |
| Z | 97 | hCoV-19/Luxembourg/LNS7802129/2020\|EPI_ISL_744633\| | ············T······· | ···················· | ···················· | ···················· | ···················· | ···················· |
| Z | 98 | hCoV-19/Luxembourg/LNS7808343/2020\|EPI_ISL_744634\| | ···················· | ···················· | ···················· | ···················· | ···················· | ···················· |
| Z | 99 | hCoV-19/Luxembourg/LNS7808479/2020\|EPI_ISL_744635\| | ···················· | ···················· | ···················· | ···················· | ···················· | ···················· |
| Z | 100 | hCoV-19/Luxembourg/LNS7809848/2020\|EPI_ISL_744636\| | ···················· | ···················· | ···················· | ···················· | ···················· | ···················· |
| Z | 101 | hCoV-19/Luxembourg/LNS7817652/2020\|EPI_ISL_744637\| | ···················· | ···················· | ···················· | ···················· | ···················· | ···················· |
| Z | 102 | hCoV-19/Luxembourg/LNS7832954/2020\|EPI_ISL_744639\| | ···················· | ···················· | ···················· | ···················· | ···················· | ···················· |
| Z | 103 | hCoV-19/Luxembourg/LNS7841987/2020\|EPI_ISL_744640\| | ···················· | ···················· | ···················· | ···················· | ···················· | ···················· |
| Z | 104 | hCoV-19/Luxembourg/LNS7844504/2020\|EPI_ISL_744641\| | ···················· | ···················· | ···················· | ···················· | ···················· | ···················· |
| Z | 105 | hCoV-19/Luxembourg/LNS7844782/2020\|EPI_ISL_744642\| | ···················· | ···················· | ···················· | ···················· | ···················· | ···················· |
| Z | 106 | hCoV-19/Luxembourg/LNS7851926/2020\|EPI_ISL_744643\| | ···················· | G··················· | ···················· | ···················· | ···················· | ···················· |
| Z | 107 | hCoV-19/Luxembourg/LNS7874511/2020\|EPI_ISL_744645\| | ···················· | ···················· | ···················· | ···················· | ···················· | ···················· |
| Z | 108 | hCoV-19/Luxembourg/LNS7878146/2020\|EPI_ISL_744646\| | ···················· | ···················· | ···················· | ···················· | ·····T·············· | ···················· |
| Z | 109 | hCoV-19/Luxembourg/LNS7891598/2020\|EPI_ISL_744648\| | ···················· | ···················· | ···················· | ···················· | ···················· | ···················· |
| Z | 110 | hCoV-19/Luxembourg/LNS7892400/2020\|EPI_ISL_744649\| | ···················· | ···················· | ···················· | ···················· | ···················· | ···················· |
| Z | 111 | hCoV-19/Luxembourg/LNS7903628/2020\|EPI_ISL_744650\| | ···················· | ···················· | ···················· | ···················· | ···················· | ···················· |
| Z | 112 | hCoV-19/Luxembourg/LNS7911820/2020\|EPI_ISL_744653\| | ···················· | ···················· | ···················· | ···················· | ···················· | ···················· |
| Z | 113 | hCoV-19/Luxembourg/LNS7915037/2020\|EPI_ISL_744654\| | ···················· | ···················· | ···················· | ···················· | ···················· | ···················· |
| Z | 114 | hCoV-19/Luxembourg/LNS7930773/2020\|EPI_ISL_744655\| | ···················· | ···················· | ···················· | ···················· | ···················· | ···················· |
| Z | 115 | hCoV-19/Luxembourg/LNS7934541/2020\|EPI_ISL_744656\| | ···················· | ···················· | ···················· | ···················· | ·····T·············· | ···················· |
| Z | 116 | hCoV-19/Luxembourg/LNS7936669/2020\|EPI_ISL_744657\| | ···················· | ···················· | ···················· | ···················· | ···················· | ···················· |
| Z | 117 | hCoV-19/Luxembourg/LNS7937572/2020\|EPI_ISL_744658\| | ···T················ | ···················· | ···················· | ···················· | ···················· | ···················· |
| Z | 118 | hCoV-19/Luxembourg/LNS7948120/2020\|EPI_ISL_744659\| | ···················· | ····A··············· | ···················· | ···················· | ···················· | ···················· |
| Z | 119 | hCoV-19/Luxembourg/LNS7948422/2020\|EPI_ISL_744660\| | ···················· | ···················· | ···················· | ···················· | ···················· | ···················· |
| Z | 120 | hCoV-19/Luxembourg/LNS7953037/2020\|EPI_ISL_744661\| | ···················· | ···················· | ···················· | ···················· | ···················· | ···················· |
| Z | 121 | hCoV-19/Luxembourg/LNS7974800/2020\|EPI_ISL_744662\| | ···················· | ···················· | ···················· | ···················· | ···················· | ···················· |
| Z | 122 | hCoV-19/Luxembourg/LNS7977901/2020\|EPI_ISL_744663\| | ···················· | ···················· | ···················· | ···················· | ···················· | ···················· |
| Z | 123 | hCoV-19/Luxembourg/LNS7983594/2020\|EPI_ISL_744664\| | ···················· | ···················· | ···················· | ···················· | ···················· | ···················· |
| Z | 124 | hCoV-19/Luxembourg/LNS7986348/2020\|EPI_ISL_744665\| | ···················· | ···················· | ···················· | ···················· | ···················· | ···················· |
| Z | 125 | hCoV-19/Luxembourg/LNS7990137/2020\|EPI_ISL_744666\| | ···················· | ···················· | ···················· | ···················· | ···················· | ···················· |
| Z | 126 | hCoV-19/Luxembourg/LNS7994655/2020\|EPI_ISL_744667\| | ···················· | ···················· | ···················· | ···················· | ···················· | ···················· |
| Z | 127 | hCoV-19/Luxembourg/LNS8005266/2020\|EPI_ISL_744668\| | ···················· | ···················· | ···················· | ···················· | ···················· | ···················· |
| Z | 128 | hCoV-19/Luxembourg/LNS8007299/2020\|EPI_ISL_744669\| | ···················· | ···················· | ···················· | ···················· | ···················· | ···················· |
| Z | 129 | hCoV-19/Luxembourg/LNS8012882/2020\|EPI_ISL_744670\| | ···················· | ···················· | ···················· | ···················· | ···················· | ···················· |
| Z | 130 | hCoV-19/Luxembourg/LNS8017005/2020\|EPI_ISL_744671\| | ···················· | ···················· | ···················· | ···················· | ···················· | ···················· |
| Z | 131 | hCoV-19/Luxembourg/LNS8019108/2020\|EPI_ISL_744672\| | ···················· | ···················· | ···················· | ···················· | ···················· | ···················· |
| Z | 132 | hCoV-19/Luxembourg/LNS8040373/2020\|EPI_ISL_744673\| | ···················· | ···················· | ···················· | ···················· | ···················· | ···················· |
| Z | 133 | hCoV-19/Luxembourg/LNS8043806/2020\|EPI_ISL_744674\| | ···················· | ···················· | ···················· | ···················· | ···················· | ···················· |
| Z | 134 | hCoV-19/Luxembourg/LNS8064090/2020\|EPI_ISL_744675\| | ···················· | ···················· | ···················· | ···················· | ···················· | ···················· |
| Z | 135 | hCoV-19/Luxembourg/LNS8070453/2020\|EPI_ISL_744676\| | ···················· | ···················· | ···················· | ···················· | ···················· | ···················· |
| Z | 136 | hCoV-19/Luxembourg/LNS8076261/2020\|EPI_ISL_744678\| | ···················· | ···················· | ···················· | ···················· | ···················· | ···················· |
| Z | 137 | hCoV-19/Luxembourg/LNS8076949/2020\|EPI_ISL_744679\| | ···················· | ···················· | ···················· | ···················· | ···················· | ···················· |
| Z | 138 | hCoV-19/Luxembourg/LNS8077542/2020\|EPI_ISL_744680\| | ···················· | ···················· | ···················· | ···················· | ···················· | ···················· |
| Z | 139 | hCoV-19/Luxembourg/LNS8092973/2020\|EPI_ISL_744681\| | ···················· | ···················· | ···················· | ···················· | ···················· | ···················· |
| Z | 140 | hCoV-19/Luxembourg/LNS8100747/2020\|EPI_ISL_744682\| | ···················· | ···················· | ···················· | ···················· | ···················· | ···················· |
| Z | 141 | hCoV-19/Luxembourg/LNS8105664/2020\|EPI_ISL_744683\| | ···················· | ···················· | ···················· | ···················· | ···················· | ···················· |
| Z | 142 | hCoV-19/Luxembourg/LNS8114560/2020\|EPI_ISL_744684\| | ···················· | ···················· | ···················· | ···················· | ···················· | ···················· |
| Z | 143 | hCoV-19/Luxembourg/LNS8117508/2020\|EPI_ISL_744685\| | ···················· | ···················· | ···················· | ···················· | ···················· | ···················· |
| Z | 144 | hCoV-19/Luxembourg/LNS8120658/2020\|EPI_ISL_744686\| | ···················· | ···················· | ···················· | ···················· | ···················· | ···················· |
| Z | 145 | hCoV-19/Luxembourg/LNS8137479/2020\|EPI_ISL_744687\| | ···················· | ···················· | ···················· | ···················· | ···················· | ···················· |
| Z | 146 | hCoV-19/Luxembourg/LNS8141064/2020\|EPI_ISL_744688\| | ···················· | ···················· | ···················· | ···················· | ···················· | ···················· |
| Z | 147 | hCoV-19/Luxembourg/LNS8152962/2020\|EPI_ISL_744689\| | ···················· | G··················· | ···················· | ···················· | ···················· | ···················· |

```
Z   148 hCoV-19/Luxembourg/LNS8157144/2020|EPI_ISL_744690|   ····················  ····················  ····················  ····················  ······T·············  ····················
Z   149 hCoV-19/Luxembourg/LNS8164067/2020|EPI_ISL_744692|   ····················  ····················  ····················  ····················  ····················  ····················
Z   150 hCoV-19/Luxembourg/LNS8164261/2020|EPI_ISL_744693|   ····················  ····················  ····················  ····················  ····················  ····················
Z   151 hCoV-19/Luxembourg/LNS8172443/2020|EPI_ISL_744694|   ····················  ····················  ····················  ····················  ····················  ····················
Z   152 hCoV-19/Luxembourg/LNS8175845/2020|EPI_ISL_744695|   ····················  ····················  ····················  ····················  ····················  ····················
Z   153 hCoV-19/Luxembourg/LNS8176465/2020|EPI_ISL_744696|   ····················  ····················  ····················  ····················  ····················  ····················
Z   154 hCoV-19/Luxembourg/LNS8177468/2020|EPI_ISL_744697|   ····················  ····················  ····················  ····················  ····················  ····················
Z   155 hCoV-19/Luxembourg/LNS8179404/2020|EPI_ISL_744698|   ····················  ····················  ····················  ····················  ····················  ····················
Z   156 hCoV-19/Luxembourg/LNS8187762/2020|EPI_ISL_744699|   ····················  ····················  ····················  ····················  ····················  ····················
Z   157 hCoV-19/USA/UT-UPHL-2012605887/2020|EPI_ISL_745900   ····················  ····················  ····················  ····················  ····················  ····················
Z   158 hCoV-19/USA/UT-UPHL-2012809254/2020|EPI_ISL_745901   ····················  ····················  ····················  ····················  ····················  ····················
Z   159 hCoV-19/USA/UT-UPHL-2012713281/2020|EPI_ISL_745902   ····················  ····················  ····················  ····················  ····················  ····················
Z   160 hCoV-19/USA/UT-UPHL-2012759080/2020|EPI_ISL_745903   ····················  ····················  ····················  ····················  ····················  ····················
Z   161 hCoV-19/USA/UT-UPHL-2012010851/2020|EPI_ISL_745904   ····················  ····················  ····················  ····················  ····················  ····················
Z   162 hCoV-19/USA/UT-UPHL-2012247385/2020|EPI_ISL_745905   ····················  ·A··················  ····················  ····················  ····················  ····················
Z   163 hCoV-19/USA/UT-UPHL-2012266527/2020|EPI_ISL_745906   ····················  ·A··················  ····················  ····················  ····················  ····················
Z   164 hCoV-19/USA/UT-UPHL-2012286179/2020|EPI_ISL_745907   ····················  ····················  ····················  ····················  ····················  ····················
Z   165 hCoV-19/USA/UT-UPHL-2012438025/2020|EPI_ISL_745908   ····················  ····················  ····················  ····················  ····················  ····················
Z   166 hCoV-19/USA/UT-UPHL-2012841191/2020|EPI_ISL_745909   ····················  ····················  ····················  ····················  ····················  ····················
Z   167 hCoV-19/USA/UT-UPHL-2012989138/2020|EPI_ISL_745910   ····················  ·A··················  ····················  ····················  ····················  ····················
Z   168 hCoV-19/USA/UT-UPHL-2012213961/2020|EPI_ISL_745911   ····················  ····················  ····················  ····················  ····················  ····················
Z   169 hCoV-19/USA/UT-UPHL-2012239501/2020|EPI_ISL_745912   ····················  ····················  ····················  ····················  ····················  ····················
Z   170 hCoV-19/USA/UT-UPHL-2012475412/2020|EPI_ISL_745913   ····················  ····················  ····················  ····················  ····················  ····················
Z   171 hCoV-19/USA/UT-UPHL-2012955404/2020|EPI_ISL_745914   ····················  ····················  ····················  ····················  ····················  ····················
Z   172 hCoV-19/USA/UT-UPHL-2012889221/2020|EPI_ISL_745915   ····················  ····················  ····················  ····················  ····················  ····················
Z   173 hCoV-19/USA/UT-UPHL-2012881074/2020|EPI_ISL_745916   ····················  ····················  ····················  ····················  ····················  ····················
Z   174 hCoV-19/USA/UT-UPHL-2012877200/2020|EPI_ISL_745917   ····················  ····················  ····················  ····················  ····················  ····················
Z   175 hCoV-19/USA/UT-UPHL-2012838775/2020|EPI_ISL_745918   ····················  ····················  ····················  ····················  ····················  ····················
Z   176 hCoV-19/USA/UT-UPHL-2012288941/2020|EPI_ISL_745919   ····················  ····················  ····················  ····················  ····················  ····················
Z   177 hCoV-19/USA/UT-UPHL-2012047870/2020|EPI_ISL_745920   ····················  ····················  ····················  ····················  ····················  ····················
Z   178 hCoV-19/USA/UT-UPHL-2012262261/2020|EPI_ISL_745921   ····················  ·A··················  ····················  ····················  ····················  ····················
Z   179 hCoV-19/USA/UT-UPHL-2012636964/2020|EPI_ISL_745922   ····················  ····················  ····················  ····················  ····················  ····················
Z   180 hCoV-19/USA/UT-UPHL-2012538290/2020|EPI_ISL_745923   ····················  ·A··················  ····················  ····················  ····················  ····················
Z   181 hCoV-19/USA/UT-UPHL-2012233218/2020|EPI_ISL_745924   ····················  ····················  ····················  ····················  ····················  ····················
Z   182 hCoV-19/USA/UT-UPHL-2012640082/2020|EPI_ISL_745925   ····················  ····················  ····················  ····················  ····················  ····················
Z   183 hCoV-19/USA/UT-UPHL-2012036591/2020|EPI_ISL_745926   ····················  ····················  ····················  ····················  ····················  ····················
Z   184 hCoV-19/USA/UT-UPHL-2012429034/2020|EPI_ISL_745927   ····················  ·A··················  ····················  ····················  ····················  ····················
Z   185 hCoV-19/USA/UT-UPHL-2012256510/2020|EPI_ISL_745928   ····················  ·A··················  ····················  ····················  ····················  ····················
Z   186 hCoV-19/USA/UT-UPHL-2012750625/2020|EPI_ISL_745929   ····················  ····················  ····················  ····················  ····················  ····················
Z   187 hCoV-19/USA/UT-UPHL-2012939669/2020|EPI_ISL_745930   ····················  ····················  ····················  ····················  ····················  ····················
Z   188 hCoV-19/USA/UT-UPHL-2012242806/2020|EPI_ISL_745931   ····················  ····················  ····················  ····················  ····················  ····················
Z   189 hCoV-19/USA/UT-UPHL-2012405767/2020|EPI_ISL_745932   ····················  ·A··················  ····················  ····················  ····················  ····················
Z   190 hCoV-19/USA/UT-UPHL-2012519953/2020|EPI_ISL_745933   ····················  ····················  ····················  ····················  ····················  ····················
Z   191 hCoV-19/USA/UT-UPHL-2012699833/2020|EPI_ISL_745934   ····················  ····················  ····················  ····················  ····················  ····················
Z   192 hCoV-19/USA/UT-UPHL-2012787067/2020|EPI_ISL_745935   ····················  ····················  ····················  ····················  ····················  ····················
Z   193 hCoV-19/USA/UT-UPHL-2012470388/2020|EPI_ISL_745936   ····················  ····················  ····················  ····················  ····················  ····················
Z   194 hCoV-19/USA/UT-UPHL-2012299429/2020|EPI_ISL_745937   ····················  ····················  ····················  ····················  ····················  ····················
Z   195 hCoV-19/USA/UT-UPHL-2012364630/2020|EPI_ISL_745938   ····················  ····················  ····················  ····················  ····················  ····················
Z   196 hCoV-19/USA/UT-UPHL-2012509614/2020|EPI_ISL_745939   ····················  ·A··················  ····················  ····················  ····················  ····················
Z   197 hCoV-19/USA/UT-UPHL-2012626021/2020|EPI_ISL_745940   ····················  ····················  ····················  ····················  ····················  ····················
Z   198 hCoV-19/USA/UT-UPHL-2012568023/2020|EPI_ISL_745941   ····················  ·A··················  ····················  ····················  ····················  ····················
Z   199 hCoV-19/USA/UT-UPHL-2012334615/2020|EPI_ISL_745942   ····················  ····················  ····················  ····················  ····················  ····················
Z   200 hCoV-19/USA/UT-UPHL-2012121783/2020|EPI_ISL_745943   ····················  ·A··················  ····················  ····················  ····················  ····················
Z   201 hCoV-19/USA/UT-UPHL-2012427828/2020|EPI_ISL_745944   ····················  ·A··················  ····················  ····················  ····················  ····················
Z   202 hCoV-19/USA/UT-UPHL-2012792358/2020|EPI_ISL_745945   ····················  ·A··················  ····················  ····················  ····················  ····················
Z   203 hCoV-19/USA/UT-UPHL-2012773576/2020|EPI_ISL_745946   ····················  ·A··················  ····················  ····················  ····················  ····················
Z   204 hCoV-19/USA/UT-UPHL-2012773500/2020|EPI_ISL_745947   ····················  ····················  ····················  ····················  ····················  ····················
Z   205 hCoV-19/USA/UT-UPHL-2012220465/2020|EPI_ISL_745948   ····················  ····················  ····················  ····················  ····················  ····················
Z   206 hCoV-19/USA/UT-UPHL-2012768090/2020|EPI_ISL_745949   ····················  ····················  ····················  ····················  ····················  ····················
Z   207 hCoV-19/USA/UT-UPHL-2012505034/2020|EPI_ISL_745950   ····················  ····················  ····················  ····················  ····················  ····················
Z   208 hCoV-19/USA/UT-UPHL-2012071943/2020|EPI_ISL_745951   ····················  ····················  ····················  ····················  ····················  ····················
Z   209 hCoV-19/USA/UT-UPHL-2012067652/2020|EPI_ISL_745952   ····················  ····················  ····················  ····················  ····················  ····················
Z   210 hCoV-19/USA/UT-UPHL-2012737021/2020|EPI_ISL_745953   ····················  ····················  ····················  ····················  ····················  ····················
Z   211 hCoV-19/USA/UT-UPHL-2012082051/2020|EPI_ISL_745954   ····················  ····················  ····················  ····················  ····················  ····················
Z   212 hCoV-19/USA/UT-UPHL-2012773632/2020|EPI_ISL_745955   ····················  ····················  ····················  ····················  ····················  ····················
Z   213 hCoV-19/USA/UT-UPHL-2012882861/2020|EPI_ISL_745956   ····················  ·A··················  ····················  ····················  ····················  ····················
Z   214 hCoV-19/USA/UT-UPHL-2012364740/2020|EPI_ISL_745957   ····················  ····················  ····················  ····················  ····················  ····················
Z   215 hCoV-19/USA/UT-UPHL-2012634496/2020|EPI_ISL_745958   ····················  ····················  ····················  ····················  ····················  ····················
Z   216 hCoV-19/USA/UT-UPHL-2012004612/2020|EPI_ISL_745959   ····················  ····················  ····················  ····················  ····················  ····················
Z   217 hCoV-19/USA/UT-UPHL-2012918171/2020|EPI_ISL_745960   ····················  ····················  ····················  ····················  ····················  ····················
Z   218 hCoV-19/USA/UT-UPHL-2012817286/2020|EPI_ISL_745961   ····················  ·A··················  ····················  ····················  ····················  ····················
Z   219 hCoV-19/USA/UT-UPHL-2012370469/2020|EPI_ISL_745962   ····················  ····················  ····················  ····················  ····················  ····················
Z   220 hCoV-19/USA/UT-UPHL-2012863994/2020|EPI_ISL_745963   ····················  ·A··················  ····················  ····················  ····················  ····················
Z   221 hCoV-19/USA/UT-UPHL-2012105116/2020|EPI_ISL_745964   ····················  ····················  ····················  ····················  ····················  ····················
Z   222 hCoV-19/USA/UT-UPHL-2012259833/2020|EPI_ISL_745965   ····················  ····················  ····················  ····················  ····················  ····················
Z   223 hCoV-19/USA/UT-UPHL-2012447003/2020|EPI_ISL_745966   ····················  ·A··················  ····················  ····················  ····················  ····················
Z   224 hCoV-19/USA/UT-UPHL-2012864126/2020|EPI_ISL_745967   ····················  ····················  ····················  ····················  ····················  ····················
Z   225 hCoV-19/USA/UT-UPHL-2012205637/2020|EPI_ISL_745968   ····················  ·A··················  ····················  ····················  ····················  ····················
Z   226 hCoV-19/USA/UT-UPHL-2012673943/2020|EPI_ISL_745969   ····················  ····················  ····················  ····················  ····················  ····················
Z   227 hCoV-19/USA/UT-UPHL-2012207652/2020|EPI_ISL_745970   ····················  ····················  ····················  ····················  ····················  ····················
```

```
Z   228 hCoV-19/USA/UT-UPHL-2012847498/2020|EPI_ISL_745971   ····················  ····················  ····················  ····················  ····················  ····················
Z   229 hCoV-19/USA/UT-UPHL-2012928787/2020|EPI_ISL_745972   ····················  ·A··················  ····················  ····················  ····················  ····················
Z   230 hCoV-19/USA/UT-UPHL-2012133597/2020|EPI_ISL_745973   ····················  ····················  ····················  ····················  ····················  ····················
Z   231 hCoV-19/USA/UT-UPHL-2012424564/2020|EPI_ISL_745974   ····················  ····················  ····················  ····················  ····················  ····················
Z   232 hCoV-19/USA/UT-UPHL-2012400155/2020|EPI_ISL_745975   ····················  ····················  ····················  ····················  ····················  ····················
Z   233 hCoV-19/USA/UT-UPHL-2012110234/2020|EPI_ISL_745976   ····················  ····················  ····················  ····················  ····················  ····················
Z   234 hCoV-19/USA/UT-UPHL-2012443791/2020|EPI_ISL_745977   ····················  ·A··················  ····················  ····················  ····················  ····················
Z   235 hCoV-19/USA/UT-UPHL-2012669236/2020|EPI_ISL_745978   ····················  ····················  ····················  ····················  ····················  ····················
Z   236 hCoV-19/USA/UT-UPHL-2012506501/2020|EPI_ISL_745979   ····················  ····················  ····················  ····················  ····················  ····················
Z   237 hCoV-19/USA/UT-UPHL-2012725139/2020|EPI_ISL_745980   ····················  ····················  ····················  ····················  ····················  ····················
Z   238 hCoV-19/USA/UT-UPHL-2012416147/2020|EPI_ISL_745981   ····················  ····················  ····················  ····················  ····················  ····················
Z   239 hCoV-19/USA/UT-UPHL-2012212287/2020|EPI_ISL_745982   ····················  ····················  ····················  ····················  ····················  ····················
Z   240 hCoV-19/USA/UT-UPHL-2012798103/2020|EPI_ISL_745983   ····················  ····················  ····················  ····················  ····················  ····················
Z   241 hCoV-19/USA/UT-UPHL-2012708431/2020|EPI_ISL_745984   ····················  ····················  ····················  ····················  ····················  ····················
Z   242 hCoV-19/USA/UT-UPHL-2012277335/2020|EPI_ISL_745985   ····················  ·A··················  ····················  ····················  ····················  ····················
Z   243 hCoV-19/USA/UT-UPHL-2012129901/2020|EPI_ISL_745986   ····················  ····················  ····················  ····················  ····················  ····················
Z   244 hCoV-19/USA/UT-UPHL-2012940320/2020|EPI_ISL_745987   ····················  ····················  ····················  ····················  ····················  ····················
Z   245 hCoV-19/USA/UT-UPHL-2012432100/2020|EPI_ISL_745988   ····················  ····················  ····················  ····················  ····················  ····················
Z   246 hCoV-19/USA/UT-UPHL-2012933851/2020|EPI_ISL_745989   ····················  ····················  ····················  ····················  ····················  ····················
Z   247 hCoV-19/USA/UT-UPHL-2012383236/2020|EPI_ISL_745990   ····················  ····················  ····················  ····················  ····················  ····················
Z   248 hCoV-19/USA/UT-UPHL-2012498286/2020|EPI_ISL_745991   ····················  ····················  ····················  ····················  ····················  ····················
Z   249 hCoV-19/USA/UT-UPHL-2012848813/2020|EPI_ISL_745992   ····················  ····················  ····················  ····················  ····················  ····················
Z   250 hCoV-19/USA/UT-UPHL-2012896199/2020|EPI_ISL_745993   ····················  ····················  ····················  ····················  ····················  ····················
Z   251 hCoV-19/USA/UT-UPHL-2012782986/2020|EPI_ISL_745994   ····················  ·A··················  ····················  ····················  ····················  ····················
Z   252 hCoV-19/USA/UT-UPHL-2012854358/2020|EPI_ISL_745995   ····················  ·A··················  ····················  ····················  ····················  ····················
Z   253 hCoV-19/USA/UT-UPHL-2012501801/2020|EPI_ISL_745996   ····················  ····················  ····················  ····················  ····················  ····················
Z   254 hCoV-19/USA/UT-UPHL-2012996508/2020|EPI_ISL_745997   ····················  ····················  ····················  ····················  ····················  ····················
Z   255 hCoV-19/USA/UT-UPHL-2012706136/2020|EPI_ISL_745998   ····················  ····················  ····················  ····················  ····················  ····················
Z   256 hCoV-19/USA/UT-UPHL-2012363851/2020|EPI_ISL_745999   ····················  ····················  ····················  ····················  ····················  ····················
Z   257 hCoV-19/USA/CA-CZB-13895/2020|EPI_ISL_738800|2020-   ····················  ····················  ····················  ····················  ····················  ····················
Z   258 hCoV-19/USA/CA-CZB-14833/2020|EPI_ISL_738801|2020-   ····················  ·A··················  ····················  ····················  ····················  ····················
Z   260 hCoV-19/USA/CA-CZB-15135/2020|EPI_ISL_738803|2020-   ····················  ·A··················  ····················  ····················  ····················  ····················
Z   261 hCoV-19/USA/CA-CZB-14059/2020|EPI_ISL_738804|2020-   ····················  ····················  ····················  ····················  ····················  ····················
Z   262 hCoV-19/USA/CA-CZB-15958/2020|EPI_ISL_738805|2020-   ····················  ·A··················  ····················  ····················  ····················  ····················
Z   263 hCoV-19/USA/CA-CZB-14802/2020|EPI_ISL_738806|2020-   ····················  ····················  ····················  ····················  ····················  ····················
Z   264 hCoV-19/USA/CA-CZB-14829/2020|EPI_ISL_738807|2020-   ····················  ····················  ····················  ····················  ····················  ····················
Z   265 hCoV-19/USA/CA-CZB-15463/2020|EPI_ISL_738808|2020-   ····················  ····················  ··T·················  ····················  ····················  ····················
Z   266 hCoV-19/USA/CA-CZB-14857/2020|EPI_ISL_738809|2020-   ····················  ····················  ····················  ····················  ····················  ····················
Z   267 hCoV-19/USA/CA-CZB-14784/2020|EPI_ISL_738810|2020-   ····················  ····················  ····················  ····················  ····················  ····················
Z   268 hCoV-19/USA/CA-CZB-14653/2020|EPI_ISL_738811|2020-   ····················  ····················  ····················  ····················  ····················  ····················
Z   272 hCoV-19/USA/CA-CZB-14228/2020|EPI_ISL_738815|2020-   ····················  ····················  ····················  ····················  ····················  ····················
Z   273 hCoV-19/USA/CA-CZB-15427/2020|EPI_ISL_738816|2020-   ····················  ····················  ····················  ····················  ····················  ····················
Z   274 hCoV-19/USA/CA-CZB-14074/2020|EPI_ISL_738817|2020-   ····················  ····················  ····················  ····················  ····················  ····················
Z   275 hCoV-19/USA/CA-CZB-14654/2020|EPI_ISL_738818|2020-   ····················  ····················  ····················  ····················  ····················  ····················
Z   277 hCoV-19/USA/CA-CZB-11254/2020|EPI_ISL_738820|2020-   ····················  ····················  ····················  ····················  ····················  ····················
Z   278 hCoV-19/USA/CA-CZB-14002/2020|EPI_ISL_738821|2020-   ····················  ····················  ····················  ····················  ····················  ····················
Z   279 hCoV-19/USA/CA-CZB-14067/2020|EPI_ISL_738822|2020-   ····················  ····················  ····················  ····················  ····················  ····················
Z   280 hCoV-19/USA/CA-CZB-14901/2020|EPI_ISL_738823|2020-   ····················  ····················  ····················  ····················  ····················  ····················
Z   281 hCoV-19/USA/CA-CZB-14645/2020|EPI_ISL_738824|2020-   ····················  ····················  ····················  ····················  ····················  ····················
Z   282 hCoV-19/USA/CA-CZB-13906/2020|EPI_ISL_738825|2020-   ····················  ·A··················  ····················  ····················  ····················  ····················
Z   283 hCoV-19/USA/CA-CZB-14887/2020|EPI_ISL_738826|2020-   ····················  ····················  ····················  ····················  ····················  ····················
Z   284 hCoV-19/USA/CA-CZB-14773/2020|EPI_ISL_738827|2020-   ····················  ····················  ····················  ····················  ····················  ····················
Z   285 hCoV-19/USA/CA-CZB-14092/2020|EPI_ISL_738828|2020-   ····················  ····················  ····················  ····················  ····················  ····················
Z   286 hCoV-19/USA/CA-CZB-15469/2020|EPI_ISL_738829|2020-   ····················  ····················  ····················  ····················  ····················  ····················
Z   287 hCoV-19/USA/CA-CZB-14678/2020|EPI_ISL_738830|2020-   ····················  ····················  ····················  ····················  ····················  ····················
Z   289 hCoV-19/USA/CA-CZB-14759/2020|EPI_ISL_738832|2020-   ····················  ····················  ····················  ····················  ····················  ····················
Z   291 hCoV-19/USA/CA-CZB-15949/2020|EPI_ISL_738834|2020-   ····················  ····················  ····················  ····················  ····················  ····················
Z   292 hCoV-19/USA/CA-CZB-13844/2020|EPI_ISL_738835|2020-   ····················  ·A··················  ····················  ····················  NNN·················  ····················   1
Z   293 hCoV-19/USA/CA-CZB-14606/2020|EPI_ISL_738836|2020-   ····················  ····················  ····················  ····················  ····················  ····················
Z   294 hCoV-19/USA/CA-CZB-14075/2020|EPI_ISL_738837|2020-   ····················  ····················  ····················  ····················  ····················  ····················
Z   295 hCoV-19/USA/CA-CZB-13946/2020|EPI_ISL_738838|2020-   ····················  ····················  ····················  ····················  ····················  ····················
Z   297 hCoV-19/USA/CA-CZB-14891/2020|EPI_ISL_738840|2020-   ····················  ····················  ····················  ····················  ····················  ····················
Z   298 hCoV-19/USA/CA-CZB-14900/2020|EPI_ISL_738841|2020-   ····················  ····················  ····················  ····················  ····················  ····················
Z   299 hCoV-19/USA/CA-CZB-15038/2020|EPI_ISL_738842|2020-   ····················  ·A··················  ····················  ····················  ····················  ····················
Z   300 hCoV-19/USA/CA-CZB-14111/2020|EPI_ISL_738843|2020-   ····················  ····················  ····················  ····················  ····················  ····················
Z   301 hCoV-19/USA/CA-CZB-14769/2020|EPI_ISL_738844|2020-   ····················  ·A··················  ····················  ····················  ····················  ····················
Z   302 hCoV-19/USA/CA-CZB-14674/2020|EPI_ISL_738845|2020-   ····················  ····················  ····················  ····················  ····················  ····················
Z   304 hCoV-19/USA/CA-CZB-14418/2020|EPI_ISL_738847|2020-   ····················  ····················  ····················  ····················  ····················  ····················
Z   305 hCoV-19/USA/CA-CZB-13848/2020|EPI_ISL_738848|2020-   ····················  ·A··················  ····················  ····················  ····················  ····················
Z   306 hCoV-19/USA/CA-CZB-14805/2020|EPI_ISL_738849|2020-   ····················  ····················  ····················  ····················  ····················  ····················
Z   307 hCoV-19/USA/CA-CZB-15473/2020|EPI_ISL_738850|2020-   ····················  ····················  ··T·················  ····················  ····················  ····················
Z   308 hCoV-19/USA/CA-CZB-16179/2020|EPI_ISL_738851|2020-   ····················  ····················  ····················  ····················  ····················  ····················
Z   309 hCoV-19/USA/CA-CZB-14771/2020|EPI_ISL_738852|2020-   ····················  ····················  ····················  ····················  ····················  ····················
Z   310 hCoV-19/USA/CA-CZB-13683/2020|EPI_ISL_738853|2020-   ····················  ·A··················  ····················  ····················  ····················  ····················
Z   311 hCoV-19/USA/CA-CZB-14850/2020|EPI_ISL_738854|2020-   ····················  ····················  ····················  ····················  ····················  ····················
Z   312 hCoV-19/USA/CA-CZB-15289/2020|EPI_ISL_738855|2020-   ····················  ····················  ····················  ····················  N···················  NNN·················   1
Z   313 hCoV-19/USA/CA-CZB-15474/2020|EPI_ISL_738856|2020-   ····················  ····················  ··T·················  ····················  ····················  ····················
Z   314 hCoV-19/USA/CA-CZB-14658/2020|EPI_ISL_738857|2020-   ····················  ·A··················  ····················  ····················  ····················  ····················
Z   315 hCoV-19/USA/CA-CZB-14136/2020|EPI_ISL_738858|2020-   ····················  ····················  ····················  ····················  ····················  ····················
Z   316 hCoV-19/USA/CA-CZB-15274/2020|EPI_ISL_738859|2020-   ····················  ····················  ····················  ····················  ····················  ····················
```

| | | | | | | | |
|---|---|---|---|---|---|---|---|
| Z | 317 hCoV-19/USA/CA-CZB-14642/2020\|EPI_ISL_738860\|2020- | ···················· | ···················· | ····················· | ···················· | ···················· | ···················· |
| Z | 319 hCoV-19/USA/CA-CZB-14710/2020\|EPI_ISL_738862\|2020- | ···················· | ···················· | ····················· | ···················· | ···················· | ···················· |
| Z | 320 hCoV-19/USA/CA-CZB-14915/2020\|EPI_ISL_738863\|2020- | ···················· | ···················· | ····················· | ···················· | ···················· | ···················· |
| Z | 321 hCoV-19/USA/CA-CZB-14993/2020\|EPI_ISL_738864\|2020- | ···················· | ·A·················· | ····················· | ···················· | ···················· | ···················· |
| Z | 322 hCoV-19/USA/CA-CZB-14825/2020\|EPI_ISL_738865\|2020- | ···················· | ···················· | ····················· | ···················· | ···················· | ···················· |
| Z | 323 hCoV-19/USA/CA-CZB-15478/2020\|EPI_ISL_738866\|2020- | ···················· | ···················· | ····················· | ···················· | ···················· | ···················· |
| Z | 324 hCoV-19/USA/CA-CZB-15037/2020\|EPI_ISL_738867\|2020- | ···················· | ·A·················· | ····················· | ···················· | ···················· | ···················· |
| Z | 325 hCoV-19/USA/CA-CZB-13737/2020\|EPI_ISL_738868\|2020- | ···················· | ·A·················· | ····················· | ···················· | ···················· | ···················· |
| Z | 326 hCoV-19/USA/CA-CZB-14238/2020\|EPI_ISL_738869\|2020- | ···················· | ···················· | ····················· | ···················· | ···················· | ···················· |
| Z | 327 hCoV-19/USA/CA-CZB-11275/2020\|EPI_ISL_738870\|2020- | ···················· | ···················· | ····················· | ···················· | ···················· | ···················· |
| Z | 328 hCoV-19/USA/CA-CZB-14236/2020\|EPI_ISL_738871\|2020- | ···················· | ···················· | ····················· | ···················· | ···················· | ···················· |
| Z | 329 hCoV-19/USA/CA-CZB-15468/2020\|EPI_ISL_738872\|2020- | ···················· | ···················· | ···T················· | ···················· | ···················· | ···················· |
| Z | 330 hCoV-19/USA/CA-CZB-14709/2020\|EPI_ISL_738873\|2020- | ···················· | ···················· | ····················· | ···················· | ···················· | ···················· |
| Z | 331 hCoV-19/USA/CA-CZB-15439/2020\|EPI_ISL_738874\|2020- | ···················· | ···················· | ····················· | ···················· | ···················· | ···················· |
| Z | 332 hCoV-19/USA/CA-CZB-14823/2020\|EPI_ISL_738875\|2020- | ···················· | ·A·················· | ····················· | ···················· | ···················· | ···················· |
| Z | 333 hCoV-19/USA/CA-CZB-14073/2020\|EPI_ISL_738876\|2020- | ···················· | ···················· | ····················· | ···················· | ···················· | ···················· |
| Z | 334 hCoV-19/USA/CA-CZB-16043/2020\|EPI_ISL_738877\|2020- | ···················· | ·A·················· | ····················· | ···················· | ···················· | ···················· |
| Z | 335 hCoV-19/USA/CA-CZB-15422/2020\|EPI_ISL_738878\|2020- | ···················· | ·A·················· | ····················· | ···················· | ···················· | ···················· |
| Z | 336 hCoV-19/USA/CA-CZB-14748/2020\|EPI_ISL_738879\|2020- | ···················· | ·A·················· | ···········T········· | ···················· | ···················· | ···················· |
| Z | 337 hCoV-19/USA/CA-CZB-15458/2020\|EPI_ISL_738880\|2020- | ···················· | ···················· | ····················· | ···················· | ···················· | ···················· |
| Z | 338 hCoV-19/USA/CA-CZB-14034/2020\|EPI_ISL_738881\|2020- | ···················· | ···················· | ····················· | ···················· | ···················· | ···················· |
| Z | 339 hCoV-19/USA/CA-CZB-14202/2020\|EPI_ISL_738882\|2020- | ···················· | ···················· | ····················· | ···················· | ···················· | ···················· |
| Z | 341 hCoV-19/USA/CA-CZB-14662/2020\|EPI_ISL_738884\|2020- | ···················· | ···················· | ····················· | ···················· | ···················· | ···················· |
| Z | 342 hCoV-19/USA/CA-CZB-14219/2020\|EPI_ISL_738885\|2020- | ···················· | ···················· | ····················· | ···················· | ···················· | ···················· |
| Z | 345 hCoV-19/USA/CA-CZB-14217/2020\|EPI_ISL_738888\|2020- | ···················· | ···················· | ····················· | ···················· | ···················· | ···················· |
| Z | 346 hCoV-19/USA/CA-CZB-15191/2020\|EPI_ISL_738889\|2020- | ···················· | ···················· | ····················· | ···················· | ···················· | ···················· |
| Z | 347 hCoV-19/USA/CA-CZB-15482/2020\|EPI_ISL_738890\|2020- | ···················· | ·A·················· | ····················· | ···················· | ···················· | ···················· |
| Z | 348 hCoV-19/USA/CA-CZB-14400/2020\|EPI_ISL_738891\|2020- | ···················· | ···················· | ····················· | ···················· | ···················· | ···················· |
| Z | 349 hCoV-19/USA/CA-CZB-14069/2020\|EPI_ISL_738892\|2020- | ···················· | ···················· | ····················· | ···················· | ···················· | ···················· |
| Z | 350 hCoV-19/USA/CA-CZB-14886/2020\|EPI_ISL_738893\|2020- | ···················· | ·A·················· | ····················· | ···················· | ···················· | ···················· |
| Z | 351 hCoV-19/USA/CA-CZB-15283/2020\|EPI_ISL_738894\|2020- | ···················· | ···················· | ····················· | ···················· | ···················· | ···················· |
| Z | 352 hCoV-19/USA/CA-CZB-14431/2020\|EPI_ISL_738895\|2020- | ···················· | ···················· | ····················· | ···················· | ···················· | ···················· |
| Z | 353 hCoV-19/USA/CA-CZB-14044/2020\|EPI_ISL_738896\|2020- | ···················· | ···················· | ····················· | ···················· | ···················· | ···················· |
| Z | 354 hCoV-19/USA/CA-CZB-11250/2020\|EPI_ISL_738897\|2020- | ···················· | ···················· | ····················· | ···················· | ···················· | ···················· |
| Z | 356 hCoV-19/USA/CA-CZB-14906/2020\|EPI_ISL_738899\|2020- | ···················· | ···················· | ····················· | ···················· | ···················· | ···················· |
| Z | 357 hCoV-19/England/SHEF-10D2E64/2020\|EPI_ISL_742051\|2 | ···················· | ···················· | ····················· | ···················· | ···················· | ···················· |
| Z | 358 hCoV-19/England/SHEF-10D3012/2020\|EPI_ISL_742053\|2 | ···················· | ···················· | ····················· | ···················· | ···················· | ···················· |
| Z | 359 hCoV-19/England/SHEF-10D374A/2020\|EPI_ISL_742059\|2 | ···················· | ···················· | ····················· | ···················· | ···················· | ···················· |
| Z | 360 hCoV-19/England/SHEF-10D3D1B/2020\|EPI_ISL_742062\|2 | ···················· | ···················· | ····················· | ···················· | ···················· | ···················· |
| Z | 361 hCoV-19/England/SHEF-10D46F1/2020\|EPI_ISL_742065\|2 | ···················· | ···················· | ····················· | ···················· | ···················· | ···················· |
| Z | 362 hCoV-19/England/SHEF-10D4A40/2020\|EPI_ISL_742068\|2 | ···················· | ···················· | ····················· | ···················· | ···················· | ···················· |
| Z | 363 hCoV-19/England/SHEF-10D5186/2020\|EPI_ISL_742072\|2 | ···················· | ···················· | ····················· | ···················· | ···················· | ···················· |
| Z | 364 hCoV-19/England/SHEF-10D5818/2020\|EPI_ISL_742078\|2 | ···················· | ···················· | ····················· | ···················· | ···················· | ···················· |
| Z | 365 hCoV-19/England/SHEF-10D5C94/2020\|EPI_ISL_742081\|2 | ···················· | ···················· | ····················· | ···················· | ···················· | ···················· |
| Z | 366 hCoV-19/England/SHEF-10D62A0/2020\|EPI_ISL_742084\|2 | ···················· | ···················· | ····················· | ···················· | ···················· | ···················· |
| Z | 367 hCoV-19/England/SHEF-10D6A99/2020\|EPI_ISL_742091\|2 | ···················· | ···················· | ····················· | ···················· | ···················· | ···················· |
| Z | 368 hCoV-19/England/SHEF-10D6D45/2020\|EPI_ISL_742094\|2 | ···················· | ···················· | ····················· | ···················· | ···················· | ···················· |
| Z | 369 hCoV-19/England/SHEF-10D70D2/2020\|EPI_ISL_742095\|2 | ···················· | ···················· | ····················· | ···················· | ···················· | ···················· |
| Z | 370 hCoV-19/England/SHEF-10D7166/2020\|EPI_ISL_742097\|2 | ···················· | ···················· | ····················· | ···················· | ···················· | ···················· |
| Z | 371 hCoV-19/England/SHEF-10D7825/2020\|EPI_ISL_742104\|2 | ···················· | ···················· | ····················· | ···················· | ···················· | ···················· |
| Z | 372 hCoV-19/England/SHEF-10D7931/2020\|EPI_ISL_742105\|2 | ···················· | ···················· | ····················· | ···················· | ···················· | ···················· |
| Z | 373 hCoV-19/England/SHEF-10D7B3B/2020\|EPI_ISL_742107\|2 | ···················· | ···················· | ····················· | ···················· | ···················· | ···················· |
| Z | 374 hCoV-19/England/SHEF-10D7E05/2020\|EPI_ISL_742110\|2 | ···················· | ···················· | ····················· | ···················· | ···················· | ···················· |
| Z | 375 hCoV-19/Northern Ireland/NIRE-23562C/2020\|EPI_ISL_ | ···················· | ···················· | ····················· | ···················· | ···················· | ···················· |
| Z | 376 hCoV-19/Northern Ireland/NIRE-23564A/2020\|EPI_ISL_ | ···················· | ···················· | ····················· | ···················· | ···················· | ···················· |
| Z | 377 hCoV-19/Northern Ireland/NIRE-235677/2020\|EPI_ISL_ | ···················· | ···················· | ····················· | ···················· | ···················· | ···················· |
| Z | 378 hCoV-19/Northern Ireland/NIRE-235686/2020\|EPI_ISL_ | ···················· | ···················· | ····················· | ···················· | ···················· | ···················· |
| Z | 379 hCoV-19/Northern Ireland/NIRE-235695/2020\|EPI_ISL_ | ···················· | ···················· | ····················· | ···················· | ···················· | ···················· |
| Z | 380 hCoV-19/Northern Ireland/NIRE-2356A4/2020\|EPI_ISL_ | ···················· | ···················· | ····················· | ···················· | ···················· | ···················· |
| Z | 381 hCoV-19/Northern Ireland/NIRE-235923/2020\|EPI_ISL_ | ···················· | ···················· | ····················· | ···················· | ···················· | ···················· |
| Z | 387 hCoV-19/Wales/PHWC-4A0086/2020\|EPI_ISL_743391\|2020 | ···················· | ···················· | ····················· | ···················· | ···················· | ···················· |
| Z | 388 hCoV-19/Wales/PHWC-4A00B3/2020\|EPI_ISL_743393\|2020 | ···················· | ···················· | ····················· | ···················· | ···················· | ···················· |
| Z | 392 hCoV-19/Wales/PHWC-4A0129/2020\|EPI_ISL_743399\|2020 | ···················· | ···················· | ····················· | ···················· | ···················· | ···················· |
| Z | 394 hCoV-19/Wales/PHWC-4A0147/2020\|EPI_ISL_743401\|2020 | ···················· | ···················· | ····················· | ···················· | ···················· | ···················· |
| Z | 395 hCoV-19/Wales/PHWC-4A0156/2020\|EPI_ISL_743402\|2020 | ···················· | ···················· | ····················· | ···················· | ···················· | ···················· |
| Z | 396 hCoV-19/Wales/PHWC-4A0165/2020\|EPI_ISL_743403\|2020 | ···················· | ···················· | ····················· | ···················· | ···················· | ···················· |
| Z | 397 hCoV-19/Wales/PHWC-4A01A1/2020\|EPI_ISL_743406\|2020 | ···················· | ···················· | ····················· | ···················· | ···················· | ···················· |
| Z | 398 hCoV-19/Wales/PHWC-4A01CF/2020\|EPI_ISL_743408\|2020 | ···················· | ···················· | ····················· | ···················· | ···················· | ···················· |
| Z | 399 hCoV-19/Wales/PHWC-4A01DE/2020\|EPI_ISL_743409\|2020 | ···················· | ···················· | ····················· | ···················· | ···················· | ···················· |
| Z | 400 hCoV-19/Wales/PHWC-4A01ED/2020\|EPI_ISL_743410\|2020 | ···················· | ···················· | ····················· | ···················· | ···················· | ···················· |
| Z | 401 hCoV-19/Wales/PHWC-4A0208/2020\|EPI_ISL_743412\|2020 | ···················· | ···················· | ····················· | ···················· | ···················· | ···················· |
| Z | 402 hCoV-19/Wales/PHWC-4A0217/2020\|EPI_ISL_743413\|2020 | ···················· | ···················· | ····················· | ···················· | ···················· | ···················· |
| Z | 403 hCoV-19/Wales/PHWC-4A0226/2020\|EPI_ISL_743414\|2020 | ···················· | ···················· | ····················· | ···················· | ···················· | ···················· |
| Z | 404 hCoV-19/Wales/PHWC-4A0244/2020\|EPI_ISL_743416\|2020 | ···················· | ···················· | ····················· | ···················· | ···················· | ···················· |
| Z | 405 hCoV-19/Wales/PHWC-4A0262/2020\|EPI_ISL_743418\|2020 | ···················· | ···················· | ····················· | ···················· | ···················· | ···················· |
| Z | 406 hCoV-19/Wales/PHWC-4A0280/2020\|EPI_ISL_743420\|2020 | ···················· | ···················· | ····················· | ···················· | ···················· | ···················· |
| Z | 407 hCoV-19/Wales/PHWC-4A029F/2020\|EPI_ISL_743421\|2020 | ···················· | ···················· | ····················· | ···················· | ···················· | ···················· |
| Z | 408 hCoV-19/Wales/PHWC-4A02AE/2020\|EPI_ISL_743422\|2020 | ···················· | ···················· | ····················· | ···················· | ···················· | ···················· |
| Z | 409 hCoV-19/Wales/PHWC-4A02EA/2020\|EPI_ISL_743426\|2020 | ···················· | ···················· | ····················· | ···················· | ···················· | ···················· |
| Z | 410 hCoV-19/Wales/PHWC-4A02F9/2020\|EPI_ISL_743427\|2020 | ···················· | ···················· | ····················· | ···················· | ···················· | ···················· |

<table>
<tr><td>Z</td><td>411</td><td>hCoV-19/Wales/PHWC-4A0323/2020|EPI_ISL_743430|2020</td><td>····················</td><td>····················</td><td>·····················</td><td>····················</td><td>····················</td><td>····················</td></tr>
<tr><td>Z</td><td>412</td><td>hCoV-19/Wales/PHWC-4A0332/2020|EPI_ISL_743431|2020</td><td>····················</td><td>····················</td><td>·····················</td><td>····················</td><td>····················</td><td>····················</td></tr>
<tr><td>Z</td><td>414</td><td>hCoV-19/Wales/PHWC-4A038D/2020|EPI_ISL_743436|2020</td><td>····················</td><td>····················</td><td>·····················</td><td>····················</td><td>····················</td><td>····················</td></tr>
<tr><td>Z</td><td>418</td><td>hCoV-19/Wales/PHWC-4A03F6/2020|EPI_ISL_743441|2020</td><td>····················</td><td>····················</td><td>·····················</td><td>····················</td><td>····················</td><td>····················</td></tr>
<tr><td>Z</td><td>419</td><td>hCoV-19/Wales/PHWC-4A0402/2020|EPI_ISL_743442|2020</td><td>····················</td><td>····················</td><td>·····················</td><td>····················</td><td>····················</td><td>····················</td></tr>
<tr><td>Z</td><td>420</td><td>hCoV-19/Wales/PHWC-4A044E/2020|EPI_ISL_743445|2020</td><td>····················</td><td>····················</td><td>·····················</td><td>····················</td><td>····················</td><td>····················</td></tr>
<tr><td>Z</td><td>421</td><td>hCoV-19/Wales/PHWC-4A046C/2020|EPI_ISL_743447|2020</td><td>····················</td><td>····················</td><td>·····················</td><td>····················</td><td>····················</td><td>····················</td></tr>
<tr><td>Z</td><td>422</td><td>hCoV-19/Wales/PHWC-4A0499/2020|EPI_ISL_743450|2020</td><td>····················</td><td>····················</td><td>·····················</td><td>····················</td><td>····················</td><td>····················</td></tr>
<tr><td>Z</td><td>423</td><td>hCoV-19/Wales/PHWC-4A04B7/2020|EPI_ISL_743452|2020</td><td>····················</td><td>····················</td><td>·····················</td><td>····················</td><td>····················</td><td>····················</td></tr>
<tr><td>Z</td><td>428</td><td>hCoV-19/Wales/PHWC-4A051E/2020|EPI_ISL_743458|2020</td><td>····················</td><td>····················</td><td>·····················</td><td>····················</td><td>····················</td><td>····················</td></tr>
<tr><td>Z</td><td>429</td><td>hCoV-19/Wales/PHWC-4A07EB/2020|EPI_ISL_743495|2020</td><td>····················</td><td>····················</td><td>·····················</td><td>····················</td><td>····················</td><td>····················</td></tr>
<tr><td>Z</td><td>430</td><td>hCoV-19/Wales/PHWC-4A0815/2020|EPI_ISL_743498|2020</td><td>····················</td><td>····················</td><td>·····················</td><td>····················</td><td>····················</td><td>····················</td></tr>
<tr><td>Z</td><td>431</td><td>hCoV-19/Luxembourg/LNS8189784/2020|EPI_ISL_744700|</td><td>····················</td><td>····················</td><td>·····················</td><td>····················</td><td>······T·············</td><td>····················</td></tr>
<tr><td>Z</td><td>432</td><td>hCoV-19/Luxembourg/LNS8194078/2020|EPI_ISL_744701|</td><td>····················</td><td>····················</td><td>·····················</td><td>····················</td><td>····················</td><td>····················</td></tr>
<tr><td>Z</td><td>433</td><td>hCoV-19/Luxembourg/LNS8198863/2020|EPI_ISL_744702|</td><td>····················</td><td>G···················</td><td>·····················</td><td>····················</td><td>····················</td><td>····················</td></tr>
<tr><td>Z</td><td>434</td><td>hCoV-19/Luxembourg/LNS8204416/2020|EPI_ISL_744703|</td><td>····················</td><td>····················</td><td>·····················</td><td>····················</td><td>····················</td><td>····················</td></tr>
<tr><td>Z</td><td>435</td><td>hCoV-19/Luxembourg/LNS8213159/2020|EPI_ISL_744704|</td><td>····················</td><td>····················</td><td>·····················</td><td>····················</td><td>····················</td><td>····················</td></tr>
<tr><td>Z</td><td>436</td><td>hCoV-19/Luxembourg/LNS8217011/2020|EPI_ISL_744705|</td><td>····················</td><td>····················</td><td>·····················</td><td>····················</td><td>····················</td><td>····················</td></tr>
<tr><td>Z</td><td>437</td><td>hCoV-19/Luxembourg/LNS8230662/2020|EPI_ISL_744706|</td><td>····················</td><td>····················</td><td>·····················</td><td>····················</td><td>····················</td><td>····················</td></tr>
<tr><td>Z</td><td>438</td><td>hCoV-19/Luxembourg/LNS8235233/2020|EPI_ISL_744707|</td><td>····················</td><td>····················</td><td>·····················</td><td>····················</td><td>····················</td><td>····················</td></tr>
<tr><td>Z</td><td>439</td><td>hCoV-19/Luxembourg/LNS8245232/2020|EPI_ISL_744708|</td><td>····················</td><td>····················</td><td>·····················</td><td>····················</td><td>····················</td><td>····················</td></tr>
<tr><td>Z</td><td>440</td><td>hCoV-19/Luxembourg/LNS8265593/2020|EPI_ISL_744711|</td><td>····················</td><td>····················</td><td>·····················</td><td>····················</td><td>····················</td><td>····················</td></tr>
<tr><td>Z</td><td>441</td><td>hCoV-19/Luxembourg/LNS8287433/2020|EPI_ISL_744712|</td><td>····················</td><td>····················</td><td>·····················</td><td>····················</td><td>····················</td><td>····················</td></tr>
<tr><td>Z</td><td>442</td><td>hCoV-19/Luxembourg/LNS8287925/2020|EPI_ISL_744713|</td><td>····················</td><td>····················</td><td>·····················</td><td>····················</td><td>····················</td><td>····················</td></tr>
<tr><td>Z</td><td>443</td><td>hCoV-19/Luxembourg/LNS8292645/2020|EPI_ISL_744714|</td><td>····················</td><td>····················</td><td>·····················</td><td>····················</td><td>····················</td><td>····················</td></tr>
<tr><td>Z</td><td>444</td><td>hCoV-19/Luxembourg/LNS8299549/2020|EPI_ISL_744715|</td><td>····················</td><td>····················</td><td>·····················</td><td>····················</td><td>····················</td><td>····················</td></tr>
<tr><td>Z</td><td>445</td><td>hCoV-19/Luxembourg/LNS8313411/2020|EPI_ISL_744716|</td><td>····················</td><td>····················</td><td>·····················</td><td>····················</td><td>····················</td><td>····················</td></tr>
<tr><td>Z</td><td>446</td><td>hCoV-19/Luxembourg/LNS8314674/2020|EPI_ISL_744717|</td><td>····················</td><td>····················</td><td>·····················</td><td>····················</td><td>····················</td><td>····················</td></tr>
<tr><td>Z</td><td>447</td><td>hCoV-19/Luxembourg/LNS8325961/2020|EPI_ISL_744718|</td><td>····················</td><td>····················</td><td>·····················</td><td>····················</td><td>····················</td><td>····················</td></tr>
<tr><td>Z</td><td>448</td><td>hCoV-19/Luxembourg/LNS8330290/2020|EPI_ISL_744719|</td><td>····················</td><td>····················</td><td>·····················</td><td>····················</td><td>····················</td><td>····················</td></tr>
<tr><td>Z</td><td>449</td><td>hCoV-19/Luxembourg/LNS8330596/2020|EPI_ISL_744720|</td><td>····················</td><td>····················</td><td>·····················</td><td>····················</td><td>····················</td><td>····················</td></tr>
<tr><td>Z</td><td>450</td><td>hCoV-19/Luxembourg/LNS8332571/2020|EPI_ISL_744721|</td><td>····················</td><td>····················</td><td>·····················</td><td>····················</td><td>····················</td><td>····················</td></tr>
<tr><td>Z</td><td>451</td><td>hCoV-19/Luxembourg/LNS8340197/2020|EPI_ISL_744722|</td><td>····················</td><td>····················</td><td>·····················</td><td>····················</td><td>····················</td><td>····················</td></tr>
<tr><td>Z</td><td>452</td><td>hCoV-19/Luxembourg/LNS8340264/2020|EPI_ISL_744723|</td><td>····················</td><td>····················</td><td>·····················</td><td>····················</td><td>····················</td><td>····················</td></tr>
<tr><td>Z</td><td>453</td><td>hCoV-19/Luxembourg/LNS8345736/2020|EPI_ISL_744724|</td><td>····················</td><td>····················</td><td>·····················</td><td>····················</td><td>····················</td><td>····················</td></tr>
<tr><td>Z</td><td>454</td><td>hCoV-19/Luxembourg/LNS8359141/2020|EPI_ISL_744726|</td><td>····················</td><td>····················</td><td>·····················</td><td>····················</td><td>····················</td><td>····················</td></tr>
<tr><td>Z</td><td>455</td><td>hCoV-19/Luxembourg/LNS8359656/2020|EPI_ISL_744727|</td><td>····················</td><td>····················</td><td>·····················</td><td>····················</td><td>····················</td><td>····················</td></tr>
<tr><td>Z</td><td>456</td><td>hCoV-19/Luxembourg/LNS8367919/2020|EPI_ISL_744728|</td><td>····················</td><td>····················</td><td>·····················</td><td>····················</td><td>····················</td><td>····················</td></tr>
<tr><td>Z</td><td>457</td><td>hCoV-19/Luxembourg/LNS8369120/2020|EPI_ISL_744729|</td><td>····················</td><td>····················</td><td>·····················</td><td>····················</td><td>····················</td><td>····················</td></tr>
<tr><td>Z</td><td>458</td><td>hCoV-19/Luxembourg/LNS8376337/2020|EPI_ISL_744730|</td><td>····················</td><td>G···················</td><td>·····················</td><td>····················</td><td>····················</td><td>····················</td></tr>
<tr><td>Z</td><td>459</td><td>hCoV-19/Luxembourg/LNS8387253/2020|EPI_ISL_744732|</td><td>····················</td><td>····················</td><td>·····················</td><td>····················</td><td>····················</td><td>····················</td></tr>
<tr><td>Z</td><td>460</td><td>hCoV-19/Luxembourg/LNS8411846/2020|EPI_ISL_744733|</td><td>····················</td><td>····················</td><td>·····················</td><td>····················</td><td>····················</td><td>····················</td></tr>
<tr><td>Z</td><td>461</td><td>hCoV-19/Luxembourg/LNS8419054/2020|EPI_ISL_744734|</td><td>····················</td><td>····················</td><td>·····················</td><td>····················</td><td>····················</td><td>····················</td></tr>
<tr><td>Z</td><td>462</td><td>hCoV-19/Luxembourg/LNS8423696/2020|EPI_ISL_744735|</td><td>····················</td><td>····················</td><td>·····················</td><td>····················</td><td>····················</td><td>····················</td></tr>
<tr><td>Z</td><td>463</td><td>hCoV-19/Luxembourg/LNS8434558/2020|EPI_ISL_744737|</td><td>····················</td><td>····················</td><td>·····················</td><td>····················</td><td>····················</td><td>····················</td></tr>
<tr><td>Z</td><td>464</td><td>hCoV-19/Luxembourg/LNS8437124/2020|EPI_ISL_744738|</td><td>····················</td><td>····················</td><td>·····················</td><td>····················</td><td>····················</td><td>····················</td></tr>
<tr><td>Z</td><td>465</td><td>hCoV-19/Luxembourg/LNS8453294/2020|EPI_ISL_744739|</td><td>····················</td><td>····················</td><td>·····················</td><td>····················</td><td>····················</td><td>····················</td></tr>
<tr><td>Z</td><td>466</td><td>hCoV-19/Luxembourg/LNS8457551/2020|EPI_ISL_744740|</td><td>····················</td><td>····················</td><td>·····················</td><td>····················</td><td>····················</td><td>····················</td></tr>
<tr><td>Z</td><td>467</td><td>hCoV-19/Luxembourg/LNS8462691/2020|EPI_ISL_744741|</td><td>····················</td><td>····················</td><td>·····················</td><td>····················</td><td>····················</td><td>····················</td></tr>
<tr><td>Z</td><td>468</td><td>hCoV-19/Luxembourg/LNS8467500/2020|EPI_ISL_744742|</td><td>····················</td><td>····················</td><td>·····················</td><td>····················</td><td>····················</td><td>····················</td></tr>
<tr><td>Z</td><td>469</td><td>hCoV-19/Luxembourg/LNS8470846/2020|EPI_ISL_744743|</td><td>····················</td><td>····················</td><td>·····················</td><td>····················</td><td>····················</td><td>····················</td></tr>
<tr><td>Z</td><td>470</td><td>hCoV-19/Luxembourg/LNS8483681/2020|EPI_ISL_744744|</td><td>····················</td><td>····················</td><td>·····················</td><td>····················</td><td>····················</td><td>····················</td></tr>
<tr><td>Z</td><td>471</td><td>hCoV-19/Luxembourg/LNS8485600/2020|EPI_ISL_744745|</td><td>····················</td><td>····················</td><td>·····················</td><td>····················</td><td>····················</td><td>····················</td></tr>
<tr><td>Z</td><td>472</td><td>hCoV-19/Luxembourg/LNS8485858/2020|EPI_ISL_744746|</td><td>····················</td><td>····················</td><td>·····················</td><td>····················</td><td>····················</td><td>····················</td></tr>
<tr><td>Z</td><td>473</td><td>hCoV-19/Luxembourg/LNS8489630/2020|EPI_ISL_744747|</td><td>····················</td><td>····················</td><td>·····················</td><td>····················</td><td>····················</td><td>····················</td></tr>
<tr><td>Z</td><td>474</td><td>hCoV-19/Luxembourg/LNS8492581/2020|EPI_ISL_744748|</td><td>····················</td><td>····················</td><td>·····················</td><td>····················</td><td>····················</td><td>····················</td></tr>
<tr><td>Z</td><td>475</td><td>hCoV-19/Luxembourg/LNS8500912/2020|EPI_ISL_744749|</td><td>····················</td><td>····················</td><td>·····················</td><td>····················</td><td>····················</td><td>····················</td></tr>
<tr><td>Z</td><td>476</td><td>hCoV-19/Luxembourg/LNS8511711/2020|EPI_ISL_744750|</td><td>····················</td><td>····················</td><td>·····················</td><td>····················</td><td>····················</td><td>····················</td></tr>
<tr><td>Z</td><td>477</td><td>hCoV-19/Luxembourg/LNS8512641/2020|EPI_ISL_744751|</td><td>····················</td><td>····················</td><td>·····················</td><td>····················</td><td>····················</td><td>····················</td></tr>
<tr><td>Z</td><td>478</td><td>hCoV-19/Luxembourg/LNS8512751/2020|EPI_ISL_744752|</td><td>····················</td><td>····················</td><td>·····················</td><td>····················</td><td>····················</td><td>····················</td></tr>
<tr><td>Z</td><td>479</td><td>hCoV-19/Luxembourg/LNS8515900/2020|EPI_ISL_744753|</td><td>····················</td><td>····················</td><td>·····················</td><td>····················</td><td>····················</td><td>····················</td></tr>
<tr><td>Z</td><td>480</td><td>hCoV-19/Luxembourg/LNS8525091/2020|EPI_ISL_744754|</td><td>····················</td><td>····················</td><td>·····················</td><td>····················</td><td>····················</td><td>····················</td></tr>
<tr><td>Z</td><td>481</td><td>hCoV-19/Luxembourg/LNS8527149/2020|EPI_ISL_744755|</td><td>····················</td><td>····················</td><td>·····················</td><td>····················</td><td>····················</td><td>····················</td></tr>
<tr><td>Z</td><td>482</td><td>hCoV-19/Luxembourg/LNS8533272/2020|EPI_ISL_744756|</td><td>····················</td><td>····················</td><td>·····················</td><td>····················</td><td>····················</td><td>····················</td></tr>
<tr><td>Z</td><td>483</td><td>hCoV-19/Luxembourg/LNS8535646/2020|EPI_ISL_744757|</td><td>····················</td><td>····················</td><td>·····················</td><td>····················</td><td>····················</td><td>····················</td></tr>
<tr><td>Z</td><td>484</td><td>hCoV-19/Luxembourg/LNS8537935/2020|EPI_ISL_744758|</td><td>····················</td><td>····················</td><td>·····················</td><td>····················</td><td>····················</td><td>····················</td></tr>
<tr><td>Z</td><td>485</td><td>hCoV-19/Luxembourg/LNS8550370/2020|EPI_ISL_744759|</td><td>····················</td><td>····················</td><td>·····················</td><td>····················</td><td>····················</td><td>····················</td></tr>
<tr><td>Z</td><td>486</td><td>hCoV-19/Luxembourg/LNS8553066/2020|EPI_ISL_744760|</td><td>····················</td><td>····················</td><td>·····················</td><td>····················</td><td>····················</td><td>····················</td></tr>
<tr><td>Z</td><td>487</td><td>hCoV-19/Luxembourg/LNS8553977/2020|EPI_ISL_744761|</td><td>····················</td><td>····················</td><td>·····················</td><td>····················</td><td>····················</td><td>····················</td></tr>
<tr><td>Z</td><td>488</td><td>hCoV-19/Luxembourg/LNS8559954/2020|EPI_ISL_744762|</td><td>····················</td><td>····················</td><td>·····················</td><td>····················</td><td>····················</td><td>····················</td></tr>
<tr><td>Z</td><td>489</td><td>hCoV-19/Luxembourg/LNS8562064/2020|EPI_ISL_744763|</td><td>············T·······</td><td>····················</td><td>·····················</td><td>····················</td><td>····················</td><td>····················</td></tr>
<tr><td>Z</td><td>490</td><td>hCoV-19/Luxembourg/LNS8570671/2020|EPI_ISL_744764|</td><td>····················</td><td>····················</td><td>·····················</td><td>····················</td><td>····················</td><td>····················</td></tr>
<tr><td>Z</td><td>491</td><td>hCoV-19/Luxembourg/LNS8571011/2020|EPI_ISL_744765|</td><td>····················</td><td>····················</td><td>·····················</td><td>····················</td><td>····················</td><td>····················</td></tr>
<tr><td>Z</td><td>492</td><td>hCoV-19/Luxembourg/LNS8577854/2020|EPI_ISL_744766|</td><td>····················</td><td>····················</td><td>·····················</td><td>····················</td><td>····················</td><td>····················</td></tr>
<tr><td>Z</td><td>493</td><td>hCoV-19/Luxembourg/LNS8585990/2020|EPI_ISL_744767|</td><td>····················</td><td>····················</td><td>·····················</td><td>····················</td><td>····················</td><td>····················</td></tr>
<tr><td>Z</td><td>494</td><td>hCoV-19/Luxembourg/LNS8592530/2020|EPI_ISL_744768|</td><td>····················</td><td>····················</td><td>·····················</td><td>····················</td><td>····················</td><td>····················</td></tr>
<tr><td>Z</td><td>495</td><td>hCoV-19/Luxembourg/LNS8593378/2020|EPI_ISL_744770|</td><td>····················</td><td>····················</td><td>·····················</td><td>····················</td><td>······T·············</td><td>····················</td></tr>
<tr><td>Z</td><td>496</td><td>hCoV-19/Luxembourg/LNS8593708/2020|EPI_ISL_744771|</td><td>····················</td><td>····················</td><td>·····················</td><td>····················</td><td>····················</td><td>····················</td></tr>
<tr><td>Z</td><td>497</td><td>hCoV-19/Luxembourg/LNS8598024/2020|EPI_ISL_744772|</td><td>····················</td><td>····················</td><td>·····················</td><td>····················</td><td>····················</td><td>····················</td></tr>
<tr><td>Z</td><td>498</td><td>hCoV-19/Luxembourg/LNS8598511/2020|EPI_ISL_744773|</td><td>····················</td><td>····················</td><td>·····················</td><td>····················</td><td>····················</td><td>····················</td></tr>
</table>

```
Z    499 hCoV-19/Luxembourg/LNS8605055/2020|EPI_ISL_744774|    ····················  ····················  ·····················  ····················  ····················  ····················
Z    500 hCoV-19/Luxembourg/LNS8606331/2020|EPI_ISL_744775|    ····················  ····················  ·····················  ····················  ······T·············  ····················
Z    501 hCoV-19/Luxembourg/LNS8615469/2020|EPI_ISL_744776|    ····················  ····················  ·····················  ····················  ····················  ····················
Z    502 hCoV-19/Luxembourg/LNS8625523/2020|EPI_ISL_744777|    ····················  ····················  ·····················  ····················  ····················  ····················
Z    503 hCoV-19/Luxembourg/LNS8628138/2020|EPI_ISL_744778|    ····················  ····················  ·····················  ····················  ····················  ····················
Z    504 hCoV-19/Luxembourg/LNS8643646/2020|EPI_ISL_744779|    ····················  ····················  ·····················  ····················  ····················  ····················
Z    505 hCoV-19/Luxembourg/LNS8646215/2020|EPI_ISL_744780|    ····················  ····················  ·····················  ····················  ····················  ····················
Z    506 hCoV-19/Luxembourg/LNS8661190/2020|EPI_ISL_744782|    ············T·······  ····················  ·····················  ····················  ····················  ····················
Z    507 hCoV-19/Luxembourg/LNS8672733/2020|EPI_ISL_744783|    ····················  ····················  ·····················  ····················  ····················  ····················
Z    508 hCoV-19/Luxembourg/LNS8675312/2020|EPI_ISL_744784|    ····················  ····················  ·····················  ····················  ····················  ····················
Z    509 hCoV-19/Luxembourg/LNS8678056/2020|EPI_ISL_744785|    ····················  ····················  ·····················  ····················  ····················  ····················
Z    510 hCoV-19/Luxembourg/LNS8681870/2020|EPI_ISL_744787|    ····················  ····················  ·····················  ····················  ····················  ····················
Z    511 hCoV-19/Luxembourg/LNS8685935/2020|EPI_ISL_744788|    ····················  ····················  ·····················  ····················  ····················  ····················
Z    512 hCoV-19/Luxembourg/LNS8690581/2020|EPI_ISL_744789|    ····················  ····················  ·····················  ····················  ····················  ····················
Z    514 hCoV-19/USA/CA-CZB-16066/2020|EPI_ISL_738901|2020-    ····················  ····················  ·····················  ····················  ····················  ····················
Z    515 hCoV-19/USA/CA-CZB-14807/2020|EPI_ISL_738902|2020-    ····················  ····················  ·····················  ····················  ····················  ····················
Z    516 hCoV-19/USA/CA-CZB-15069/2020|EPI_ISL_738903|2020-    ····················  ····················  ·····················  ····················  ····················  ····················
Z    518 hCoV-19/USA/CA-CZB-14661/2020|EPI_ISL_738905|2020-    ····················  ····················  ·····················  ····················  ····················  ····················
Z    520 hCoV-19/USA/CA-CZB-14913/2020|EPI_ISL_738907|2020-    ····················  ·A··················  ·····················  ····················  ····················  ····················
Z    521 hCoV-19/USA/CA-CZB-14890/2020|EPI_ISL_738908|2020-    ····················  ····················  ·····················  ····················  ····················  ····················
Z    522 hCoV-19/USA/CA-CZB-15081/2020|EPI_ISL_738909|2020-    ····················  ····················  ·····················  ····················  ····················  ····················
Z    523 hCoV-19/USA/CA-CZB-13677/2020|EPI_ISL_738910|2020-    ····················  ·A··················  ·····················  ····················  ····················  ····················
Z    524 hCoV-19/USA/CA-CZB-11263/2020|EPI_ISL_738911|2020-    ····················  ····················  ·····················  ····················  ····················  ····················
Z    525 hCoV-19/USA/CA-CZB-14778/2020|EPI_ISL_738912|2020-    ····················  ····················  ·····················  ····················  ····················  ····················
Z    526 hCoV-19/USA/CA-CZB-14785/2020|EPI_ISL_738913|2020-    ····················  ····················  ·····················  ····················  ····················  ····················
Z    527 hCoV-19/USA/CA-CZB-13997/2020|EPI_ISL_738914|2020-    ····················  ····················  ·····················  ····················  ····················  ····················
Z    528 hCoV-19/USA/CA-CZB-15239/2020|EPI_ISL_738915|2020-    ····················  ·A··················  ·····················  ····················  ····················  ····················
Z    530 hCoV-19/USA/CA-CZB-14815/2020|EPI_ISL_738917|2020-    ····················  ····················  ·····················  ····················  ····················  ····················
Z    531 hCoV-19/USA/CA-CZB-14765/2020|EPI_ISL_738918|2020-    ····················  ····················  ·····················  ····················  ····················  ····················
Z    532 hCoV-19/USA/CA-CZB-13893/2020|EPI_ISL_738919|2020-    ····················  ····················  ·····················  ····················  ····················  ····················
Z    533 hCoV-19/USA/CA-CZB-14555/2020|EPI_ISL_738920|2020-    ····················  ····················  ·····················  ····················  ····················  ····················
Z    534 hCoV-19/USA/CA-CZB-14700/2020|EPI_ISL_738921|2020-    ····················  ····················  ·····················  ····················  ····················  ····················
Z    535 hCoV-19/USA/CA-CZB-13686/2020|EPI_ISL_738922|2020-    ····················  ····················  ·····················  ····················  ····················  ····················
Z    536 hCoV-19/USA/CA-CZB-14791/2020|EPI_ISL_738923|2020-    ····················  ····················  ·····················  ····················  ····················  ····················
Z    537 hCoV-19/USA/CA-CZB-14956/2020|EPI_ISL_738924|2020-    ····················  ·A··················  ·····················  ····················  ····················  ··················NN    1
Z    538 hCoV-19/USA/CA-CZB-14422/2020|EPI_ISL_738925|2020-    ····················  ····················  ·····················  ····················  ····················  ····················
Z    541 hCoV-19/USA/CA-CZB-14912/2020|EPI_ISL_738928|2020-    ····················  ·A··················  ·····················  ····················  ····················  ····················
Z    542 hCoV-19/USA/CA-CZB-14847/2020|EPI_ISL_738929|2020-    ····················  ····················  ·····················  ····················  ····················  ····················
Z    543 hCoV-19/USA/CA-CZB-14211/2020|EPI_ISL_738930|2020-    ····················  ·············A······  ·····················  ····················  ····················  ····················
Z    544 hCoV-19/USA/CA-CZB-15303/2020|EPI_ISL_738931|2020-    ····················  ·A··················  ·····················  ····················  ····················  ····················
Z    545 hCoV-19/USA/CA-CZB-13746/2020|EPI_ISL_738932|2020-    ····················  ·A··················  ·····················  ····················  ····················  ····················
Z    547 hCoV-19/USA/CA-CZB-13981/2020|EPI_ISL_738934|2020-    ····················  ····················  ·····················  ····················  ····················  ····················
Z    548 hCoV-19/USA/CA-CZB-14680/2020|EPI_ISL_738935|2020-    ····················  ····················  ·····················  ····················  ····················  ····················
Z    549 hCoV-19/USA/CA-CZB-15063/2020|EPI_ISL_738936|2020-    ····················  ····················  ·····················  ····················  ····················  ····················
Z    550 hCoV-19/USA/CA-CZB-14223/2020|EPI_ISL_738937|2020-    ····················  ····················  ·····················  ····················  ····················  ····················
Z    551 hCoV-19/USA/CA-CZB-14828/2020|EPI_ISL_738938|2020-    ····················  ····················  ·····················  ····················  ····················  ····················
Z    552 hCoV-19/USA/CA-CZB-15443/2020|EPI_ISL_738939|2020-    ····················  ····················  ·····················  ····················  ····················  ····················
Z    554 hCoV-19/USA/CA-CZB-15300/2020|EPI_ISL_738941|2020-    ····················  ·A··················  ·····················  ····················  ····················  ····················
Z    555 hCoV-19/USA/CA-CZB-15097/2020|EPI_ISL_738942|2020-    ····················  ····················  ·····················  ····················  ····················  ····················
Z    556 hCoV-19/USA/CA-CZB-14562/2020|EPI_ISL_738943|2020-    ····················  ····················  ·····················  ····················  ····················  ····················
Z    557 hCoV-19/USA/CA-CZB-14691/2020|EPI_ISL_738944|2020-    ····················  ····················  ·····················  ····················  ····················  ····················
Z    558 hCoV-19/USA/CA-CZB-13989/2020|EPI_ISL_738945|2020-    ····················  ····················  ·····················  ····················  ····················  ····················
Z    559 hCoV-19/USA/CA-CZB-14215/2020|EPI_ISL_738946|2020-    ····················  ····················  ·····················  ····················  ····················  ····················
Z    560 hCoV-19/USA/CA-CZB-14754/2020|EPI_ISL_738947|2020-    ····················  ····················  ·····················  ····················  ····················  ····················
Z    561 hCoV-19/USA/CA-CZB-15136/2020|EPI_ISL_738948|2020-    ····················  ·A··················  ·····················  ····················  ····················  ····················
Z    562 hCoV-19/USA/CA-CZB-15429/2020|EPI_ISL_738949|2020-    ····················  ·A··················  ·····················  ····················  ····················  ····················
Z    563 hCoV-19/USA/CA-CZB-14103/2020|EPI_ISL_738950|2020-    ····················  ····················  ·····················  ····················  ····················  ····················
Z    564 hCoV-19/USA/CA-CZB-14751/2020|EPI_ISL_738951|2020-    ····················  ····················  ·····················  ····················  ····················  ····················
Z    565 hCoV-19/USA/CA-CZB-11268/2020|EPI_ISL_738952|2020-    ····················  ····················  ·····················  ····················  ····················  ····················
Z    566 hCoV-19/USA/CA-CZB-14758/2020|EPI_ISL_738953|2020-    ····················  ·A··················  ·····················  ····················  ····················  ····················
Z    567 hCoV-19/USA/CA-CZB-15447/2020|EPI_ISL_738954|2020-    ····················  ····················  ·····················  ····················  ····················  ····················
Z    568 hCoV-19/USA/CA-CZB-15946/2020|EPI_ISL_738955|2020-    ····················  ····················  ·····················  ····················  ····················  ····················
Z    569 hCoV-19/USA/CA-CZB-13684/2020|EPI_ISL_738956|2020-    ···T················  ·A··················  ·····················  ····················  ····················  ····················
Z    571 hCoV-19/USA/CA-CZB-14855/2020|EPI_ISL_738958|2020-    ····················  ·A··················  ·····················  ····················  ····················  ····················
Z    572 hCoV-19/USA/CA-CZB-11264/2020|EPI_ISL_738959|2020-    ····················  ····················  ·····················  ····················  ····················  ····················
Z    574 hCoV-19/USA/CA-CZB-14742/2020|EPI_ISL_738961|2020-    ····················  ·A··················  ···········T·········  ····················  ····················  ····················
Z    575 hCoV-19/USA/CA-CZB-14553/2020|EPI_ISL_738962|2020-    ····················  ····················  ·····················  ····················  ····················  ····················
Z    577 hCoV-19/USA/CA-CZB-14152/2020|EPI_ISL_738964|2020-    ····················  ·A··················  ·····················  ····················  ····················  ····················
Z    578 hCoV-19/USA/CA-CZB-15068/2020|EPI_ISL_738965|2020-    ····················  ····················  ·····················  ····················  ····················  ····················
Z    580 hCoV-19/USA/CA-CZB-15943/2020|EPI_ISL_738967|2020-    ····················  ·A··················  ·····················  ····················  ····················  ····················
Z    581 hCoV-19/USA/CA-CZB-14660/2020|EPI_ISL_738968|2020-    ····················  ····················  ·····················  ····················  ····················  ····················
Z    582 hCoV-19/USA/CA-CZB-14836/2020|EPI_ISL_738969|2020-    ····················  ····················  ·····················  ····················  ····················  ····················
Z    583 hCoV-19/USA/CA-CZB-14671/2020|EPI_ISL_738970|2020-    ····················  ····················  ·····················  ····················  ····················  ····················
Z    584 hCoV-19/USA/CA-CZB-15150/2020|EPI_ISL_738971|2020-    ····················  ·A··················  ·····················  ····················  ····················  ····················
Z    585 hCoV-19/USA/CA-CZB-14528/2020|EPI_ISL_738972|2020-    ····················  ·A··················  ·····················  ····················  ····················  ····················
Z    586 hCoV-19/USA/CA-CZB-14673/2020|EPI_ISL_738973|2020-    ····················  ····················  ·····················  ····················  ····················  ····················
Z    588 hCoV-19/USA/CA-CZB-14632/2020|EPI_ISL_738975|2020-    ····················  ····················  ·····················  ····················  ····················  ····················
Z    590 hCoV-19/USA/CA-CZB-16138/2020|EPI_ISL_738977|2020-    ····················  ····················  ·····················  ····················  ····················  ····················
Z    591 hCoV-19/USA/CA-CZB-15403/2020|EPI_ISL_738978|2020-    ····················  ····················  ·····················  ····················  ····················  ····················
Z    592 hCoV-19/USA/CA-CZB-15326/2020|EPI_ISL_738979|2020-    ····················  ·A··················  ·····················  ····················  ····················  ····················
```

| | | | | | | | |
|---|---|---|---|---|---|---|---|
| Z | 593 hCoV-19/USA/CA-CZB-14824/2020\|EPI_ISL_738980\|2020- | ···················· | ···················· | ···················· | ···················· | ···················· | ···················· |
| Z | 594 hCoV-19/USA/CA-CZB-15430/2020\|EPI_ISL_738981\|2020- | ···················· | ·A·················· | ···················· | ···················· | ···················· | ···················· |
| Z | 596 hCoV-19/USA/CA-CZB-14354/2020\|EPI_ISL_738983\|2020- | ···················· | ···················· | ···················· | ···················· | ···················· | ···················· |
| Z | 597 hCoV-19/USA/CA-CZB-15298/2020\|EPI_ISL_738984\|2020- | ···················· | ·A·················· | ···················· | ···················· | ···················· | ···················· |
| Z | 598 hCoV-19/USA/CA-CZB-14782/2020\|EPI_ISL_738985\|2020- | ···················· | ···················· | ···················· | ···················· | ···················· | ···················· |
| Z | 599 hCoV-19/USA/CA-CZB-14120/2020\|EPI_ISL_738986\|2020- | ···················· | ···················· | ···················· | ···················· | ···················· | ···················· |
| Z | 600 hCoV-19/USA/CA-CZB-15155/2020\|EPI_ISL_738987\|2020- | ···················· | ·A·················· | ···················· | ···················· | ···················· | ···················· |
| Z | 601 hCoV-19/USA/CA-CZB-14734/2020\|EPI_ISL_738988\|2020- | ···················· | ·A·················· | ···········T········ | ···················· | ···················· | ···················· |
| Z | 602 hCoV-19/USA/CA-CZB-15471/2020\|EPI_ISL_738989\|2020- | ···················· | ···················· | ··T················· | ···················· | ···················· | ···················· |
| Z | 603 hCoV-19/USA/CA-CZB-15491/2020\|EPI_ISL_738990\|2020- | ···················· | ···················· | ···················· | ···················· | ···················· | ···················· |
| Z | 604 hCoV-19/USA/CA-CZB-14878/2020\|EPI_ISL_738991\|2020- | ···················· | ·A·················· | ···················· | ···················· | ···················· | ···················· |
| Z | 605 hCoV-19/USA/CA-CZB-14048/2020\|EPI_ISL_738992\|2020- | ···················· | ···················· | ···················· | ···················· | ···················· | ···················· |
| Z | 606 hCoV-19/USA/CA-CZB-15457/2020\|EPI_ISL_738993\|2020- | ···················· | ···················· | ···················· | ···················· | ···················· | ···················· |
| Z | 607 hCoV-19/USA/CA-CZB-15329/2020\|EPI_ISL_738994\|2020- | ···················· | ·A·················· | ···················· | ···················· | ···················· | ···················· |
| Z | 608 hCoV-19/USA/CA-CZB-14947/2020\|EPI_ISL_738995\|2020- | ···················· | ···················· | ···················· | ···················· | ···················· | ···················· |
| Z | 610 hCoV-19/USA/CA-CZB-15944/2020\|EPI_ISL_738997\|2020- | ···················· | ···················· | ···················· | ···················· | ···················· | ···················· |
| Z | 611 hCoV-19/USA/CA-CZB-14558/2020\|EPI_ISL_738998\|2020- | ···················· | ···················· | ···················· | ···················· | ···················· | ···················· |
| Z | 612 hCoV-19/USA/CA-CZB-14849/2020\|EPI_ISL_738999\|2020- | ···················· | ···················· | ···················· | ···················· | ···················· | ···················· |
| Z | 613 hCoV-19/Northern Ireland/NIRE-2356C2/2020\|EPI_ISL_ | ···················· | ···················· | ···················· | ···················· | ···················· | ···················· |
| Z | 614 hCoV-19/Northern Ireland/NIRE-2356D1/2020\|EPI_ISL_ | ···················· | ···················· | ···················· | ···················· | ···················· | ···················· |
| Z | 615 hCoV-19/Northern Ireland/NIRE-2356FF/2020\|EPI_ISL_ | ···················· | ···················· | ···················· | ···················· | ···················· | ···················· |
| Z | 616 hCoV-19/Northern Ireland/NIRE-23570B/2020\|EPI_ISL_ | ···················· | ···················· | ···················· | ···················· | ···················· | ···················· |
| Z | 617 hCoV-19/Northern Ireland/NIRE-23571A/2020\|EPI_ISL_ | ···················· | ···················· | ···················· | ···················· | ···················· | ···················· |
| Z | 618 hCoV-19/Northern Ireland/NIRE-235729/2020\|EPI_ISL_ | ···················· | ···················· | ···················· | ···················· | ···················· | ···················· |
| Z | 619 hCoV-19/Northern Ireland/NIRE-235738/2020\|EPI_ISL_ | ···················· | ···················· | ···················· | ···················· | ···················· | ···················· |
| Z | 620 hCoV-19/Northern Ireland/NIRE-235765/2020\|EPI_ISL_ | ···················· | ···················· | ···················· | ···················· | ···················· | ···················· |
| Z | 621 hCoV-19/Northern Ireland/NIRE-2357B0/2020\|EPI_ISL_ | ···················· | ···················· | ···················· | ···················· | ···················· | ···················· |
| Z | 622 hCoV-19/Northern Ireland/NIRE-2357CF/2020\|EPI_ISL_ | ···················· | ···················· | ···················· | ···················· | ···················· | ···················· |
| Z | 623 hCoV-19/Northern Ireland/NIRE-2357DE/2020\|EPI_ISL_ | ···················· | ···················· | ···················· | ···················· | ···················· | ···················· |
| Z | 624 hCoV-19/Northern Ireland/NIRE-2357ED/2020\|EPI_ISL_ | ···················· | ···················· | ···················· | ···················· | ···················· | ···················· |
| Z | 625 hCoV-19/Northern Ireland/NIRE-235808/2020\|EPI_ISL_ | ···················· | ···················· | ···················· | ···················· | ···················· | ···················· |
| Z | 626 hCoV-19/Northern Ireland/NIRE-235817/2020\|EPI_ISL_ | ···················· | ···················· | ···················· | ···················· | ···················· | ···················· |
| Z | 627 hCoV-19/Northern Ireland/NIRE-235826/2020\|EPI_ISL_ | ···················· | ···················· | ···················· | ···················· | ···················· | ···················· |
| Z | 628 hCoV-19/Northern Ireland/NIRE-235835/2020\|EPI_ISL_ | ···················· | ···················· | ···················· | ···················· | ···················· | ···················· |
| Z | 629 hCoV-19/Northern Ireland/NIRE-235844/2020\|EPI_ISL_ | ···················· | ···················· | ···················· | ···················· | ···················· | ···················· |
| Z | 630 hCoV-19/Northern Ireland/NIRE-235853/2020\|EPI_ISL_ | ···················· | ···················· | ···················· | ···················· | ···················· | ···················· |
| Z | 631 hCoV-19/Northern Ireland/NIRE-235862/2020\|EPI_ISL_ | ···················· | ···················· | ···················· | ···················· | ···················· | ···················· |
| Z | 632 hCoV-19/Northern Ireland/NIRE-235871/2020\|EPI_ISL_ | ···················· | ···················· | ···················· | ···················· | ···················· | ···················· |
| Z | 633 hCoV-19/Northern Ireland/NIRE-235880/2020\|EPI_ISL_ | ···················· | ···················· | ···················· | ···················· | ···················· | ···················· |
| Z | 634 hCoV-19/Northern Ireland/NIRE-23589F/2020\|EPI_ISL_ | ···················· | ···················· | ···················· | ···················· | ···················· | ···················· |
| Z | 635 hCoV-19/Northern Ireland/NIRE-2358AE/2020\|EPI_ISL_ | ···················· | ···················· | ···················· | ···················· | ···················· | ···················· |
| Z | 636 hCoV-19/Northern Ireland/NIRE-2358BD/2020\|EPI_ISL_ | ···················· | ···················· | ···················· | ···················· | ···················· | ···················· |
| Z | 637 hCoV-19/Northern Ireland/NIRE-2358EA/2020\|EPI_ISL_ | ···················· | ···················· | ···················· | ···················· | ···················· | ···················· |
| Z | 638 hCoV-19/Northern Ireland/NIRE-2358F9/2020\|EPI_ISL_ | ···················· | ···················· | ···················· | ···················· | ···················· | ···················· |
| Z | 639 hCoV-19/Northern Ireland/NIRE-23598D/2020\|EPI_ISL_ | ···················· | ···················· | ···················· | ···················· | ···················· | ···················· |
| Z | 640 hCoV-19/Northern Ireland/NIRE-23599C/2020\|EPI_ISL_ | ···················· | ···················· | ···················· | ···················· | ···················· | ···················· |
| Z | 642 hCoV-19/Scotland/EDB10891/2020\|EPI_ISL_742178\|2020 | ···················· | ···················· | ···················· | ···················· | ···················· | ···················· |
| Z | 643 hCoV-19/Scotland/EDB10907/2020\|EPI_ISL_742182\|2020 | ···················· | ···················· | ···················· | ···················· | ···················· | ···················· |
| Z | 645 hCoV-19/Scotland/EDB10915/2020\|EPI_ISL_742184\|2020 | ···················· | ···················· | ···················· | ···················· | ···················· | ···················· |
| Z | 646 hCoV-19/Scotland/EDB10922/2020\|EPI_ISL_742185\|2020 | ···················· | ···················· | ···················· | ···················· | ···················· | ···················· |
| Z | 647 hCoV-19/Scotland/EDB10938/2020\|EPI_ISL_742186\|2020 | ···················· | ···················· | ···················· | ···················· | ···················· | ···················· |
| Z | 648 hCoV-19/Scotland/EDB11000/2020\|EPI_ISL_742188\|2020 | ···················· | ···················· | ···················· | ···················· | ···················· | ···················· |
| Z | 649 hCoV-19/Scotland/EDB11006/2020\|EPI_ISL_742189\|2020 | ···················· | ···················· | ···················· | ···················· | ···················· | ···················· |
| Z | 650 hCoV-19/Scotland/EDB11011/2020\|EPI_ISL_742190\|2020 | ···················· | ···················· | ···················· | ···················· | ···················· | ···················· |
| Z | 651 hCoV-19/Scotland/EDB11012/2020\|EPI_ISL_742191\|2020 | ···················· | ···················· | ···················· | ···················· | ···················· | ···················· |
| Z | 652 hCoV-19/Scotland/EDB11013/2020\|EPI_ISL_742192\|2020 | ···················· | ···················· | ···················· | ···················· | ···················· | ···················· |
| Z | 653 hCoV-19/Scotland/EDB11026/2020\|EPI_ISL_742193\|2020 | ···················· | ···················· | ···················· | ···················· | ···················· | ···················· |
| Z | 654 hCoV-19/Scotland/EDB11035/2020\|EPI_ISL_742195\|2020 | ···················· | ···················· | ···················· | ···················· | ···················· | ···················· |
| Z | 655 hCoV-19/Scotland/EDB11046/2020\|EPI_ISL_742196\|2020 | ···················· | ···················· | ···················· | ···················· | ···················· | ···················· |
| Z | 656 hCoV-19/Scotland/EDB11055/2020\|EPI_ISL_742199\|2020 | ···················· | ···················· | ···················· | ···················· | ···················· | ···················· |
| Z | 657 hCoV-19/Scotland/EDB11074/2020\|EPI_ISL_742200\|2020 | ···················· | ···················· | ···················· | ···················· | ······T············· | ···················· |
| Z | 658 hCoV-19/Scotland/EDB11077/2020\|EPI_ISL_742201\|2020 | ···················· | ···················· | ···················· | ···················· | ···················· | ···················· |
| Z | 661 hCoV-19/Wales/PHWC-4A054B/2020\|EPI_ISL_743461\|2020 | ···················· | ···················· | ···················· | ···················· | ···················· | ···················· |
| Z | 664 hCoV-19/Wales/PHWC-4A0596/2020\|EPI_ISL_743464\|2020 | ···················· | ···················· | ···················· | ···················· | ···················· | ···················· |
| Z | 665 hCoV-19/Wales/PHWC-4A05A5/2020\|EPI_ISL_743465\|2020 | ···················· | ···················· | ···················· | ···················· | ···················· | ···················· |
| Z | 667 hCoV-19/Wales/PHWC-4A05F0/2020\|EPI_ISL_743469\|2020 | ···················· | ···················· | ···················· | ···················· | ···················· | ···················· |
| Z | 670 hCoV-19/Wales/PHWC-4A0648/2020\|EPI_ISL_743473\|2020 | ···················· | ···················· | ···················· | ···················· | ···················· | ···················· |
| Z | 671 hCoV-19/Wales/PHWC-4A0657/2020\|EPI_ISL_743474\|2020 | ···················· | ···················· | ···················· | ···················· | ···················· | ···················· |
| Z | 675 hCoV-19/Wales/PHWC-4A06DF/2020\|EPI_ISL_743482\|2020 | ···················· | ···················· | ···················· | ···················· | ···················· | ···················· |
| Z | 676 hCoV-19/Wales/PHWC-4A06FD/2020\|EPI_ISL_743484\|2020 | ···················· | ···················· | ···················· | ···················· | ···················· | ···················· |
| Z | 680 hCoV-19/Wales/PHWC-4A0745/2020\|EPI_ISL_743488\|2020 | ···················· | ···················· | ···················· | ···················· | ···················· | ···················· |
| Z | 681 hCoV-19/Wales/PHWC-4A0754/2020\|EPI_ISL_743489\|2020 | ···················· | ···················· | ···················· | ···················· | ···················· | ···················· |
| Z | 684 hCoV-19/Wales/PHWC-4A0806/2020\|EPI_ISL_743497\|2020 | ···················· | ···················· | ···················· | ···················· | ···················· | ···················· |
| Z | 685 hCoV-19/Wales/PHWC-4A098B/2020\|EPI_ISL_743520\|2020 | ···················· | ···················· | ···················· | ···················· | ···················· | ···················· |
| Z | 686 hCoV-19/Wales/PHWC-4A09D6/2020\|EPI_ISL_743525\|2020 | ···················· | ···················· | ···················· | ···················· | ···················· | ···················· |
| Z | 688 hCoV-19/Wales/PHWC-4A09F4/2020\|EPI_ISL_743527\|2020 | ···················· | ···················· | ···················· | ···················· | ···················· | ···················· |
| Z | 689 hCoV-19/Wales/PHWC-4A0A1F/2020\|EPI_ISL_743529\|2020 | ···················· | ···················· | ···················· | ···················· | ···················· | ···················· |
| Z | 690 hCoV-19/Wales/PHWC-4A0A2E/2020\|EPI_ISL_743530\|2020 | ···················· | ···················· | ···················· | ···················· | ···················· | ···················· |
| Z | 691 hCoV-19/Wales/PHWC-4A0A3D/2020\|EPI_ISL_743531\|2020 | ···················· | ···················· | ···················· | ···················· | ···················· | ···················· |
| Z | 692 hCoV-19/Wales/PHWC-4A0A4C/2020\|EPI_ISL_743532\|2020 | ···················· | ···················· | ···················· | ···················· | ···················· | ···················· |

Z 693 hCoV-19/Wales/PHWC-4A0A5B/2020|EPI_ISL_743533|2020 ···················· ···················· ···················· ···················· ···················· ····················
Z 694 hCoV-19/Wales/PHWC-4A0A6A/2020|EPI_ISL_743534|2020 ···················· ···················· ···················· ···················· ···················· ····················
Z 695 hCoV-19/Luxembourg/LNS8690605/2020|EPI_ISL_744790| ···················· ···················· ···················· ···················· ···················· ····················
Z 696 hCoV-19/Luxembourg/LNS8695282/2020|EPI_ISL_744792| ···················· ···················· ···················· ···················· ···················· ····················
Z 697 hCoV-19/Luxembourg/LNS8697915/2020|EPI_ISL_744793| ···················· ···················· ··········T········· ···················· ···················· ····················
Z 698 hCoV-19/Luxembourg/LNS8701673/2020|EPI_ISL_744794| ···················· ···················· ···················· ···················· ···················· ····················
Z 699 hCoV-19/Luxembourg/LNS8710489/2020|EPI_ISL_744795| ···················· ···················· ···················· ···················· ···················· ····················
Z 700 hCoV-19/Luxembourg/LNS8711862/2020|EPI_ISL_744796| ···················· ···················· ···················· ···················· ···················· ····················
Z 701 hCoV-19/Luxembourg/LNS8715972/2020|EPI_ISL_744797| ···················· ···················· ···················· ···················· ···················· ····················
Z 702 hCoV-19/Luxembourg/LNS8726298/2020|EPI_ISL_744798| ···················· ···················· ···················· ···················· ···················· ····················
Z 703 hCoV-19/Luxembourg/LNS8727286/2020|EPI_ISL_744799| ···················· ···················· ···················· ···················· ···················· ····················
Z 704 hCoV-19/Luxembourg/LNS8734909/2020|EPI_ISL_744802| ···················· ···················· ···················· ···················· ···················· ····················
Z 705 hCoV-19/Luxembourg/LNS8736041/2020|EPI_ISL_744803| ···················· ···················· ···················· ···················· ···················· ····················
Z 706 hCoV-19/Luxembourg/LNS8737808/2020|EPI_ISL_744804| ···················· ···················· ···················· ···················· ···················· ····················
Z 707 hCoV-19/Luxembourg/LNS8743587/2020|EPI_ISL_744805| ···················· ···················· ···················· ···················· ···················· ····················
Z 708 hCoV-19/Luxembourg/LNS8756036/2020|EPI_ISL_744806| ··········T········· ···················· ···················· ···················· ···················· ····················
Z 709 hCoV-19/Luxembourg/LNS8778022/2020|EPI_ISL_744807| ···················· ···················· ··········T········· ···················· ···················· ····················
Z 710 hCoV-19/Luxembourg/LNS8789322/2020|EPI_ISL_744808| ···················· ···················· ···················· ···················· ···················· ····················
Z 711 hCoV-19/Luxembourg/LNS8797468/2020|EPI_ISL_744809| ···················· ···················· ···················· ···················· ···················· ····················
Z 712 hCoV-19/Luxembourg/LNS8803242/2020|EPI_ISL_744810| ···················· ···················· ···················· ···················· ···················· ····················
Z 713 hCoV-19/Luxembourg/LNS8803466/2020|EPI_ISL_744811| ···················· ···················· ···················· ···················· ···················· ····················
Z 714 hCoV-19/Luxembourg/LNS8812582/2020|EPI_ISL_744812| ···················· ···················· ···················· ···················· ···················· ····················
Z 715 hCoV-19/Luxembourg/LNS8814346/2020|EPI_ISL_744813| ···················· ···················· ···················· ···················· ···················· ····················
Z 716 hCoV-19/Luxembourg/LNS8832723/2020|EPI_ISL_744814| ···················· ···················· ···················· ···················· ···················· ····················
Z 717 hCoV-19/Luxembourg/LNS8857986/2020|EPI_ISL_744815| ···················· ···················· ···················· ···················· ···················· ····················
Z 718 hCoV-19/Luxembourg/LNS8858813/2020|EPI_ISL_744816| ···················· ···················· ···················· ···················· ···················· ····················
Z 719 hCoV-19/Luxembourg/LNS8868148/2020|EPI_ISL_744817| ···················· ···················· ···················· ···················· ···················· ····················
Z 720 hCoV-19/Luxembourg/LNS8875749/2020|EPI_ISL_744818| ···················· ···················· ···················· ···················· ···················· ····················
Z 721 hCoV-19/Luxembourg/LNS8881447/2020|EPI_ISL_744819| ···················· ···················· ···················· ···················· ···················· ····················
Z 722 hCoV-19/Luxembourg/LNS8882011/2020|EPI_ISL_744820| ···················· ···················· ···················· ···················· ···················· ····················
Z 723 hCoV-19/Luxembourg/LNS8910032/2020|EPI_ISL_744821| ···················· ···················· ···················· ···················· ···················· ····················
Z 724 hCoV-19/Luxembourg/LNS8911052/2020|EPI_ISL_744822| ···················· ···················· ···················· ···················· ···················· ····················
Z 725 hCoV-19/Luxembourg/LNS9097516/2020|EPI_ISL_744850| ···················· ···················· ···················· ···················· ···················· ····················
Z 726 hCoV-19/Luxembourg/LNS9098276/2020|EPI_ISL_744851| ···················· ···················· ···················· ···················· ·····T·············· ····················
Z 727 hCoV-19/Luxembourg/LNS9120473/2020|EPI_ISL_744856| ···················· ···················· ···················· ···················· ···················· ····················
Z 728 hCoV-19/Luxembourg/LNS9132258/2020|EPI_ISL_744857| ···················· ···················· ···················· ···················· ···················· ····················
Z 729 hCoV-19/Luxembourg/LNS9142329/2020|EPI_ISL_744858| ···················· ···················· ···················· ···················· ···················· ····················
Z 730 hCoV-19/Luxembourg/LNS9145063/2020|EPI_ISL_744859| ···················· ···················· ···················· ···················· ···················· ····················
Z 731 hCoV-19/Luxembourg/LNS9146043/2020|EPI_ISL_744860| ···················· ···················· ···················· ···················· ···················· ····················
Z 732 hCoV-19/Luxembourg/LNS9151332/2020|EPI_ISL_744861| ···················· ···················· ···················· ···················· ·····T·············· ····················
Z 733 hCoV-19/Luxembourg/LNS9162230/2020|EPI_ISL_744862| ···················· ···················· ···················· ···················· ···················· ····················
Z 734 hCoV-19/Luxembourg/LNS9177314/2020|EPI_ISL_744864| ···················· ···················· ···················· ···················· ···················· ····················
Z 735 hCoV-19/Luxembourg/LNS9181195/2020|EPI_ISL_744865| ···················· ···················· ···················· ···················· ···················· ····················
Z 736 hCoV-19/Luxembourg/LNS9183553/2020|EPI_ISL_744866| ···················· ···················· ···················· ···················· ···················· ····················
Z 737 hCoV-19/USA/UT-UPHL-2012984128/2020|EPI_ISL_747000 ···················· ·A·················· ···················· ···················· ···················· ····················
Z 738 hCoV-19/USA/UT-UPHL-2012637932/2020|EPI_ISL_747001 ···················· ·A·················· ···················· ···················· ···················· ····················
Z 739 hCoV-19/USA/UT-UPHL-2012501214/2020|EPI_ISL_747002 ···················· ···················· ···················· ···················· ···················· ····················
Z 740 hCoV-19/USA/UT-UPHL-2012993930/2020|EPI_ISL_747003 ···················· ···················· ···················· ···················· ···················· ····················
Z 741 hCoV-19/USA/UT-UPHL-2012061031/2020|EPI_ISL_747004 ···················· ···················· ···················· ···················· ···················· ····················
Z 742 hCoV-19/USA/UT-UPHL-2012602372/2020|EPI_ISL_747005 ···················· ·A·················· ···················· ···················· ···················· ····················
Z 743 hCoV-19/USA/UT-UPHL-2012523456/2020|EPI_ISL_747006 ···················· ···················· ···················· ···················· ···················· ····················
Z 744 hCoV-19/USA/UT-UPHL-2012401484/2020|EPI_ISL_747007 ···················· ···················· ···················· ···················· ···················· ····················
Z 745 hCoV-19/USA/UT-UPHL-2012910509/2020|EPI_ISL_747008 ···················· ···················· ···················· ···················· ···················· ····················
Z 746 hCoV-19/USA/UT-UPHL-2012307754/2020|EPI_ISL_747009 ···················· ···················· ···················· ···················· ···················· ····················
Z 747 hCoV-19/USA/UT-UPHL-2012055574/2020|EPI_ISL_747010 ···················· ·A·················· ···················· ···················· ···················· ····················
Z 748 hCoV-19/USA/UT-UPHL-2012795207/2020|EPI_ISL_747011 ···················· ·A·················· ···················· ···················· ···················· ····················
Z 749 hCoV-19/USA/UT-UPHL-2012086088/2020|EPI_ISL_747012 ···················· ·A·················· ···················· ···················· ···················· ····················
Z 750 hCoV-19/USA/UT-UPHL-2012865596/2020|EPI_ISL_747013 ···················· ·A·················· ···················· ···················· ···················· ····················
Z 751 hCoV-19/USA/UT-UPHL-2012472233/2020|EPI_ISL_747014 ···················· ···················· ···················· ···················· ···················· ····················
Z 752 hCoV-19/USA/UT-UPHL-2012878128/2020|EPI_ISL_747015 ···················· ···················· ···················· ···················· ···················· ····················
Z 753 hCoV-19/USA/UT-UPHL-2012619954/2020|EPI_ISL_747016 ···················· ···················· ···················· ···················· ···················· ····················
Z 754 hCoV-19/USA/UT-UPHL-2012989729/2020|EPI_ISL_747017 ···················· ···················· ···················· ···················· ···················· ····················
Z 755 hCoV-19/USA/UT-UPHL-2012927532/2020|EPI_ISL_747018 ···················· ·A·················· ···················· ···················· ···················· ····················
Z 756 hCoV-19/USA/UT-UPHL-2012454470/2020|EPI_ISL_747019 ···················· ·A·················· ···················· ···················· ···················· ····················
Z 757 hCoV-19/USA/UT-UPHL-2012333679/2020|EPI_ISL_747020 ···················· ···················· ···················· ···················· ···················· ····················
Z 758 hCoV-19/USA/UT-UPHL-2012789017/2020|EPI_ISL_747021 ···················· ···················· ···················· ···················· ···················· ····················
Z 759 hCoV-19/USA/UT-UPHL-2012342729/2020|EPI_ISL_747022 ···················· ·A·················· ···················· ···················· ···················· ····················
Z 760 hCoV-19/USA/UT-UPHL-2012583894/2020|EPI_ISL_747023 ···················· ·A·················· ···················· ···················· ···················· ····················
Z 761 hCoV-19/USA/UT-UPHL-2012468949/2020|EPI_ISL_747024 ···················· ···················· ···················· ···················· ···················· ····················
Z 762 hCoV-19/USA/UT-UPHL-2012162581/2020|EPI_ISL_747025 ···················· ···················· ··T················· ···················· ···················· ····················
Z 763 hCoV-19/USA/UT-UPHL-2012888899/2020|EPI_ISL_747026 ···················· ···················· ···················· ···················· ···················· ····················
Z 764 hCoV-19/USA/UT-UPHL-2012157829/2020|EPI_ISL_747027 ···················· ·A·················· ···················· ···················· ···················· ··A·················
Z 765 hCoV-19/USA/UT-UPHL-2012469356/2020|EPI_ISL_747028 ···················· ···················· ···················· ···················· ···················· ····················
Z 766 hCoV-19/USA/UT-UPHL-2012012900/2020|EPI_ISL_747029 ···················· ···················· ···················· ···················· ···················· ····················
Z 767 hCoV-19/USA/UT-UPHL-2012527816/2020|EPI_ISL_747030 ···················· ···················· ···················· ···················· ···················· ····················
Z 768 hCoV-19/USA/UT-UPHL-2012787576/2020|EPI_ISL_747031 ·······T············ ·A·················· ···················· ···················· ···················· ····················
Z 769 hCoV-19/Scotland/EDB11115/2020|EPI_ISL_742211|2020 ···················· ···················· ···················· ···················· ···················· ····················
Z 770 hCoV-19/Scotland/EDB9555/2020|EPI_ISL_742243|2020- ···················· ···················· ···················· ···················· ···················· ····················
Z 771 hCoV-19/Scotland/EDB9762/2020|EPI_ISL_742244|2020- ···················· ···················· ···················· ···················· ···················· ····················
Z 772 hCoV-19/Scotland/EDB9885/2020|EPI_ISL_742245|2020- ···················· ···················· ···················· ···················· ···················· ····················

| | | | | | | | | |
|---|---|---|---|---|---|---|---|---|
| Z | 773 | hCoV-19/Scotland/EDB9886/2020\|EPI_ISL_742246\|2020- | ···················· | ···················· | ···················· | ···················· | ···················· | ···················· |
| Z | 774 | hCoV-19/Scotland/QEUH-B7CDA3/2020\|EPI_ISL_742247\|2 | ···················· | ···················· | ···················· | ···················· | ···················· | ···················· |
| Z | 775 | hCoV-19/Scotland/QEUH-B7CDB2/2020\|EPI_ISL_742248\|2 | ···················· | ···················· | ···················· | ···················· | ···················· | ···················· |
| Z | 776 | hCoV-19/Scotland/QEUH-B7D7A4/2020\|EPI_ISL_742249\|2 | ···················· | ···················· | ···················· | ···················· | ···················· | ···················· |
| Z | 777 | hCoV-19/Scotland/QEUH-B7D80B/2020\|EPI_ISL_742250\|2 | ···················· | ···················· | ···················· | ···················· | ···················· | ···················· |
| Z | 778 | hCoV-19/Scotland/QEUH-B7DDC0/2020\|EPI_ISL_742251\|2 | ···················· | ···················· | ···················· | ···················· | ···················· | ···················· |
| Z | 779 | hCoV-19/Scotland/QEUH-B7DE09/2020\|EPI_ISL_742252\|2 | ···················· | ···················· | ···················· | ···················· | ···················· | ···················· |
| Z | 780 | hCoV-19/Scotland/QEUH-B7DE54/2020\|EPI_ISL_742253\|2 | ···················· | ···················· | ···················· | ···················· | ···················· | ···················· |
| Z | 781 | hCoV-19/Scotland/QEUH-B7DEBE/2020\|EPI_ISL_742254\|2 | ···················· | ···················· | ···················· | ···················· | ···················· | ···················· |
| Z | 782 | hCoV-19/Scotland/QEUH-B7DEDC/2020\|EPI_ISL_742255\|2 | ···················· | ···················· | ···················· | ···················· | ···················· | ···················· |
| Z | 783 | hCoV-19/Scotland/QEUH-B7DF51/2020\|EPI_ISL_742256\|2 | ···················· | ···················· | ···················· | ···················· | ···················· | ···················· |
| Z | 784 | hCoV-19/Scotland/QEUH-B7DFAC/2020\|EPI_ISL_742257\|2 | ···················· | ···················· | ···················· | ···················· | ···················· | ···················· |
| Z | 785 | hCoV-19/Scotland/QEUH-B7E002/2020\|EPI_ISL_742258\|2 | ···················· | ···················· | ···················· | ···················· | ···················· | ···················· |
| Z | 786 | hCoV-19/Wales/ALDP-B75B82/2020\|EPI_ISL_742259\|2020 | ···················· | ···················· | ···················· | ···················· | ···················· | ···················· |
| Z | 788 | hCoV-19/Wales/PHWC-49AE76/2020\|EPI_ISL_742263\|2020 | ···················· | ···················· | ···················· | ···················· | ···················· | ···················· |
| Z | 790 | hCoV-19/Wales/PHWC-49AE94/2020\|EPI_ISL_742265\|2020 | ···················· | ···················· | ···················· | ···················· | ···················· | ···················· |
| Z | 791 | hCoV-19/Wales/PHWC-49AEA3/2020\|EPI_ISL_742266\|2020 | ···················· | ···················· | ···················· | ···················· | ···················· | ···················· |
| Z | 794 | hCoV-19/Wales/PHWC-49AED0/2020\|EPI_ISL_742269\|2020 | ···················· | ···················· | ···················· | ···················· | ···················· | ···················· |
| Z | 795 | hCoV-19/Wales/PHWC-49AEEF/2020\|EPI_ISL_742270\|2020 | ···················· | ···················· | ···················· | ···················· | ···················· | ···················· |
| Z | 797 | hCoV-19/Wales/PHWC-49AF64/2020\|EPI_ISL_742276\|2020 | ···················· | ···················· | ···················· | ···················· | ···················· | ···················· |
| Z | 798 | hCoV-19/Wales/PHWC-49AF82/2020\|EPI_ISL_742277\|2020 | ···················· | ···················· | ···················· | ···················· | ···················· | ···················· |
| Z | 799 | hCoV-19/Wales/PHWC-49AFA0/2020\|EPI_ISL_742278\|2020 | ···················· | ···················· | ···················· | ···················· | ···················· | ···················· |
| Z | 800 | hCoV-19/Wales/PHWC-49AFBF/2020\|EPI_ISL_742279\|2020 | ···················· | ···················· | ···················· | ···················· | ···················· | ···················· |
| Z | 801 | hCoV-19/Wales/PHWC-49AFEC/2020\|EPI_ISL_742281\|2020 | ···················· | ···················· | ···················· | ···················· | ···················· | ···················· |
| Z | 802 | hCoV-19/Wales/PHWC-49B006/2020\|EPI_ISL_742282\|2020 | ···················· | ···················· | ···················· | ···················· | ···················· | ···················· |
| Z | 803 | hCoV-19/Wales/PHWC-49B015/2020\|EPI_ISL_742283\|2020 | ···················· | ···················· | ···················· | ···················· | ···················· | ···················· |
| Z | 805 | hCoV-19/Wales/PHWC-49B08E/2020\|EPI_ISL_742289\|2020 | ···················· | ···················· | ···················· | ···················· | ···················· | ···················· |
| Z | 806 | hCoV-19/Wales/PHWC-49B0AC/2020\|EPI_ISL_742291\|2020 | ···················· | ···················· | ···················· | ···················· | ···················· | ···················· |
| Z | 808 | hCoV-19/Wales/PHWC-49B0CA/2020\|EPI_ISL_742293\|2020 | ···················· | ···················· | ···················· | ···················· | ···················· | ···················· |
| Z | 813 | hCoV-19/Wales/PHWC-49B112/2020\|EPI_ISL_742298\|2020 | ···················· | ···················· | ···················· | ···················· | ···················· | ···················· |
| Z | 815 | hCoV-19/Wales/PHWC-49B25B/2020\|EPI_ISL_742310\|2020 | ···················· | ···················· | ···················· | ···················· | ···················· | ···················· |
| Z | 816 | hCoV-19/Wales/PHWC-49B279/2020\|EPI_ISL_742312\|2020 | ···················· | ···················· | ···················· | ···················· | ···················· | ···················· |
| Z | 817 | hCoV-19/Wales/PHWC-49B2A6/2020\|EPI_ISL_742315\|2020 | ···················· | ···················· | ···················· | ···················· | ···················· | ···················· |
| Z | 818 | hCoV-19/Wales/PHWC-49B2B5/2020\|EPI_ISL_742316\|2020 | ···················· | ···················· | ···················· | ···················· | ···················· | ···················· |
| Z | 819 | hCoV-19/Wales/PHWC-49B2D3/2020\|EPI_ISL_742317\|2020 | ···················· | ···················· | ···················· | ···················· | ···················· | ···················· |
| Z | 820 | hCoV-19/Wales/PHWC-49B2E2/2020\|EPI_ISL_742318\|2020 | ···················· | ···················· | ···················· | ···················· | ···················· | ···················· |
| Z | 821 | hCoV-19/Wales/PHWC-49B31C/2020\|EPI_ISL_742319\|2020 | ···················· | ···················· | ···················· | ···················· | ···················· | ···················· |
| Z | 822 | hCoV-19/Wales/PHWC-49B32B/2020\|EPI_ISL_742320\|2020 | ···················· | ···················· | ···················· | ···················· | ···················· | ···················· |
| Z | 823 | hCoV-19/Wales/PHWC-49B33A/2020\|EPI_ISL_742321\|2020 | ···················· | ···················· | ···················· | ···················· | ···················· | ···················· |
| Z | 824 | hCoV-19/Wales/PHWC-49B349/2020\|EPI_ISL_742322\|2020 | ···················· | ···················· | ···················· | ···················· | ···················· | ···················· |
| Z | 825 | hCoV-19/Wales/PHWC-49B358/2020\|EPI_ISL_742323\|2020 | ···················· | ···················· | ···················· | ···················· | ···················· | ···················· |
| Z | 826 | hCoV-19/Wales/PHWC-49B376/2020\|EPI_ISL_742324\|2020 | ···················· | ···················· | ···················· | ···················· | ···················· | ···················· |
| Z | 827 | hCoV-19/Wales/PHWC-49B385/2020\|EPI_ISL_742325\|2020 | ···················· | ···················· | ···················· | ···················· | ···················· | ···················· |
| Z | 829 | hCoV-19/Wales/PHWC-4A087F/2020\|EPI_ISL_743504\|2020 | ···················· | ···················· | ···················· | ···················· | ···················· | ···················· |
| Z | 830 | hCoV-19/Wales/PHWC-4A089D/2020\|EPI_ISL_743506\|2020 | ···················· | ···················· | ···················· | ···················· | ···················· | ···················· |
| Z | 831 | hCoV-19/Wales/PHWC-4A08AC/2020\|EPI_ISL_743507\|2020 | ···················· | ···················· | ···················· | ··············A····· | ···················· | ···················· |
| Z | 833 | hCoV-19/Wales/PHWC-4A08E8/2020\|EPI_ISL_743511\|2020 | ···················· | ···················· | ···················· | ···················· | ···················· | ···················· |
| Z | 834 | hCoV-19/Wales/PHWC-4A08F7/2020\|EPI_ISL_743512\|2020 | ···················· | ···················· | ···················· | ···················· | ···················· | ···················· |
| Z | 838 | hCoV-19/Wales/PHWC-4A095E/2020\|EPI_ISL_743517\|2020 | ···················· | ···················· | ···················· | ···················· | ···················· | ···················· |
| Z | 839 | hCoV-19/Wales/PHWC-4A096D/2020\|EPI_ISL_743518\|2020 | ···················· | ···················· | ···················· | ···················· | ···················· | ···················· |
| Z | 840 | hCoV-19/Wales/PHWC-4A097C/2020\|EPI_ISL_743519\|2020 | ···················· | ···················· | ···················· | ···················· | ···················· | ···················· |
| Z | 841 | hCoV-19/Wales/PHWC-4A09A9/2020\|EPI_ISL_743522\|2020 | ···················· | ···················· | ···················· | ···················· | ···················· | ···················· |
| Z | 843 | hCoV-19/Wales/PHWC-4A0A88/2020\|EPI_ISL_743536\|2020 | ···················· | ···················· | ···················· | ···················· | ···················· | ···················· |
| Z | 844 | hCoV-19/Wales/PHWC-4A0A97/2020\|EPI_ISL_743537\|2020 | ···················· | ···················· | ···················· | ···················· | ···················· | ···················· |
| Z | 845 | hCoV-19/Wales/PHWC-4A0AA6/2020\|EPI_ISL_743538\|2020 | ···················· | ···················· | ···················· | ···················· | ···················· | ···················· |
| Z | 846 | hCoV-19/Wales/PHWC-4A0AB5/2020\|EPI_ISL_743539\|2020 | ···················· | ···················· | ···················· | ···················· | ···················· | ···················· |
| Z | 847 | hCoV-19/Wales/PHWC-4A0AC4/2020\|EPI_ISL_743540\|2020 | ···················· | ···················· | ···················· | ···················· | ···················· | ···················· |
| Z | 848 | hCoV-19/Wales/PHWC-4A0AD3/2020\|EPI_ISL_743541\|2020 | ···················· | ···················· | ···················· | ···················· | ···················· | ···················· |
| Z | 849 | hCoV-19/Wales/PHWC-4A0AE2/2020\|EPI_ISL_743542\|2020 | ···················· | ···················· | ···················· | ···················· | ···················· | ···················· |
| Z | 850 | hCoV-19/Wales/PHWC-4A0AF1/2020\|EPI_ISL_743543\|2020 | ···················· | ···················· | ···················· | ···················· | ···················· | ···················· |
| Z | 851 | hCoV-19/Wales/PHWC-4A0B1C/2020\|EPI_ISL_743545\|2020 | ···················· | ···················· | ···················· | ···················· | ···················· | ···················· |
| Z | 852 | hCoV-19/Wales/PHWC-4A0B2B/2020\|EPI_ISL_743546\|2020 | ···················· | ···················· | ···················· | ···················· | ···················· | ···················· |
| Z | 853 | hCoV-19/Wales/PHWC-4A0B3A/2020\|EPI_ISL_743547\|2020 | ···················· | ···················· | ···················· | ···················· | ···················· | ···················· |
| Z | 854 | hCoV-19/Wales/PHWC-4A0B58/2020\|EPI_ISL_743549\|2020 | ···················· | ···················· | ···················· | ···················· | ···················· | ···················· |
| Z | 855 | hCoV-19/Wales/PHWC-4A0B67/2020\|EPI_ISL_743550\|2020 | ···················· | ···················· | ···················· | ···················· | ···················· | ···················· |
| Z | 856 | hCoV-19/Wales/PHWC-4A0B76/2020\|EPI_ISL_743551\|2020 | ···················· | ···················· | ···················· | ···················· | ···················· | ···················· |
| Z | 857 | hCoV-19/Wales/PHWC-4A0B85/2020\|EPI_ISL_743552\|2020 | ···················· | ···················· | ···················· | ···················· | ···················· | ···················· |
| Z | 858 | hCoV-19/Wales/PHWC-4A0B94/2020\|EPI_ISL_743553\|2020 | ···················· | ···················· | ···················· | ···················· | ···················· | ···················· |
| Z | 859 | hCoV-19/Wales/PHWC-4A0BA3/2020\|EPI_ISL_743554\|2020 | ···················· | ···················· | ···················· | ···················· | ···················· | ···················· |
| Z | 860 | hCoV-19/Wales/PHWC-4A0BB2/2020\|EPI_ISL_743555\|2020 | ···················· | ···················· | ···················· | ···················· | ···················· | ···················· |
| Z | 861 | hCoV-19/Wales/PHWC-4A0BC1/2020\|EPI_ISL_743556\|2020 | ···················· | ···················· | ···················· | ···················· | ···················· | ···················· |
| Z | 862 | hCoV-19/Wales/PHWC-4A0BD0/2020\|EPI_ISL_743557\|2020 | ···················· | ···················· | ···················· | ···················· | ···················· | ···················· |
| Z | 863 | hCoV-19/Wales/PHWC-4A0C0A/2020\|EPI_ISL_743559\|2020 | ···················· | ···················· | ···················· | ···················· | ···················· | ···················· |
| Z | 864 | hCoV-19/Wales/PHWC-4A0C19/2020\|EPI_ISL_743560\|2020 | ···················· | ···················· | ···················· | ···················· | ···················· | ···················· |
| Z | 865 | hCoV-19/Wales/PHWC-4A0C28/2020\|EPI_ISL_743561\|2020 | ···················· | ···················· | ···················· | ···················· | ···················· | ···················· |
| Z | 866 | hCoV-19/Wales/PHWC-4A0C37/2020\|EPI_ISL_743562\|2020 | ···················· | ···················· | ···················· | ···················· | ···················· | ···················· |
| Z | 867 | hCoV-19/Wales/PHWC-4A0C46/2020\|EPI_ISL_743563\|2020 | ···················· | ···················· | ···················· | ···················· | ···················· | ···················· |
| Z | 868 | hCoV-19/Wales/PHWC-4A0C55/2020\|EPI_ISL_743564\|2020 | ···················· | ···················· | ···················· | ···················· | ···················· | ···················· |
| Z | 869 | hCoV-19/Wales/PHWC-4A0C73/2020\|EPI_ISL_743565\|2020 | ···················· | ···················· | ···················· | ···················· | ···················· | ···················· |
| Z | 870 | hCoV-19/Wales/PHWC-4A0C91/2020\|EPI_ISL_743566\|2020 | ···················· | ···················· | ···················· | ···················· | ···················· | ···················· |

| | | | | | | | |
|---|---|---|---|---|---|---|---|
| Z | 871 hCoV-19/Wales/PHWC-4A0CFB/2020\|EPI_ISL_743567\|2020 | ···················· | ···················· | ····················· | ···················· | ···················· | ···················· |
| Z | 874 hCoV-19/Wales/PHWC-4A0D43/2020\|EPI_ISL_743570\|2020 | ···················· | ···················· | ····················· | ···················· | ···················· | ···················· |
| Z | 875 hCoV-19/Wales/PHWC-4A0D52/2020\|EPI_ISL_743571\|2020 | ···················· | ···················· | ····················· | ···················· | ···················· | ···················· |
| Z | 876 hCoV-19/Wales/PHWC-4A0D70/2020\|EPI_ISL_743573\|2020 | ···················· | ···················· | ····················· | ···················· | ···················· | ···················· |
| Z | 878 hCoV-19/Wales/PHWC-4A0D9E/2020\|EPI_ISL_743575\|2020 | ···················· | ···················· | ····················· | ···················· | ···················· | ···················· |
| Z | 880 hCoV-19/Wales/PHWC-4A0DBC/2020\|EPI_ISL_743577\|2020 | ···················· | ···················· | ····················· | ···················· | ···················· | ···················· |
| Z | 881 hCoV-19/Wales/PHWC-4A0DDA/2020\|EPI_ISL_743579\|2020 | ···················· | ···················· | ····················· | ···················· | ···················· | ···················· |
| Z | 883 hCoV-19/Wales/PHWC-4A0E04/2020\|EPI_ISL_743582\|2020 | ···················· | ···················· | ····················· | ···················· | ···················· | ···················· |
| Z | 884 hCoV-19/Wales/PHWC-4A0E5F/2020\|EPI_ISL_743584\|2020 | ···················· | ···················· | ····················· | ···················· | ···················· | ···················· |
| Z | 886 hCoV-19/Wales/PHWC-4A0EAA/2020\|EPI_ISL_743588\|2020 | ···················· | ···················· | ····················· | ···················· | ···················· | ···················· |
| Z | 887 hCoV-19/Wales/PHWC-4A0EB9/2020\|EPI_ISL_743589\|2020 | ···················· | ···················· | ····················· | ···················· | ···················· | ···················· |
| Z | 888 hCoV-19/Wales/PHWC-4A0EC8/2020\|EPI_ISL_743590\|2020 | ···················· | ···················· | ····················· | ···················· | ···················· | ···················· |
| Z | 889 hCoV-19/Wales/PHWC-4A0ED7/2020\|EPI_ISL_743591\|2020 | ···················· | ···················· | ····················· | ···················· | ···················· | ···················· |
| Z | 890 hCoV-19/Wales/PHWC-4A0F2F/2020\|EPI_ISL_743596\|2020 | ···················· | ···················· | ····················· | ···················· | ···················· | ···················· |
| Z | 891 hCoV-19/Wales/PHWC-4A0F4D/2020\|EPI_ISL_743597\|2020 | ···················· | ···················· | ····················· | ···················· | ···················· | ···················· |
| Z | 892 hCoV-19/Wales/PHWC-4A0F5C/2020\|EPI_ISL_743598\|2020 | ···················· | ···················· | ····················· | ···················· | ···················· | ···················· |
| Z | 893 hCoV-19/Wales/PHWC-4A0F6B/2020\|EPI_ISL_743599\|2020 | ···················· | ···················· | ····················· | ···················· | ···················· | ···················· |
| Z | 894 hCoV-19/Wales/PHWC-4A0F7A/2020\|EPI_ISL_743600\|2020 | ···················· | ···················· | ····················· | ···················· | ···················· | ···················· |
| Z | 895 hCoV-19/Wales/PHWC-4A0F89/2020\|EPI_ISL_743601\|2020 | ···················· | ···················· | ····················· | ···················· | ···················· | ···················· |
| Z | 896 hCoV-19/Wales/PHWC-4A0F98/2020\|EPI_ISL_743602\|2020 | ···················· | ···················· | ····················· | ···················· | ···················· | ···················· |
| Z | 897 hCoV-19/Wales/PHWC-4A0FA7/2020\|EPI_ISL_743603\|2020 | ···················· | ···················· | ····················· | ···················· | ···················· | ···················· |
| Z | 899 hCoV-19/Wales/PHWC-4A1225/2020\|EPI_ISL_743640\|2020 | ···················· | ···················· | ····················· | ···················· | ···················· | ···················· |
| Z | 904 hCoV-19/Wales/PHWC-4A12BC/2020\|EPI_ISL_743649\|2020 | ···················· | ···················· | ····················· | ···················· | ···················· | ···················· |
| Z | 905 hCoV-19/Wales/PHWC-4A12CB/2020\|EPI_ISL_743650\|2020 | ···················· | ···················· | ····················· | ···················· | ···················· | ···················· |
| Z | 906 hCoV-19/Wales/PHWC-4A12DA/2020\|EPI_ISL_743651\|2020 | ···················· | ···················· | ····················· | ···················· | ···················· | ···················· |
| Z | 907 hCoV-19/Wales/PHWC-4A12E9/2020\|EPI_ISL_743652\|2020 | ···················· | ···················· | ····················· | ···················· | ···················· | ···················· |
| Z | 909 hCoV-19/Wales/PHWC-4A1304/2020\|EPI_ISL_743654\|2020 | ···················· | ···················· | ····················· | ···················· | ···················· | ···················· |
| Z | 910 hCoV-19/Wales/PHWC-4A1313/2020\|EPI_ISL_743655\|2020 | ···················· | ···················· | ····················· | ···················· | ···················· | ···················· |
| Z | 911 hCoV-19/Wales/PHWC-4A1322/2020\|EPI_ISL_743656\|2020 | ···················· | ···················· | ····················· | ···················· | ···················· | ···················· |
| Z | 912 hCoV-19/Luxembourg/LNS8924752/2020\|EPI_ISL_744823\| | ···················· | ···················· | ····················· | ···················· | ···················· | ···················· |
| Z | 913 hCoV-19/Luxembourg/LNS8929027/2020\|EPI_ISL_744824\| | ···················· | ···················· | ····················· | ···················· | ···················· | ···················· |
| Z | 914 hCoV-19/Luxembourg/LNS8945808/2020\|EPI_ISL_744825\| | ···················· | ···················· | ····················· | ···················· | ···················· | ···················· |
| Z | 915 hCoV-19/Luxembourg/LNS8948626/2020\|EPI_ISL_744826\| | ···················· | ···················· | ····················· | ···················· | ···················· | ···················· |
| Z | 916 hCoV-19/Luxembourg/LNS8959275/2020\|EPI_ISL_744827\| | ···················· | ···················· | ····················· | ···················· | ···················· | ···················· |
| Z | 917 hCoV-19/Luxembourg/LNS8965091/2020\|EPI_ISL_744828\| | ···················· | ···················· | ····················· | ···················· | ···················· | ···················· |
| Z | 918 hCoV-19/Luxembourg/LNS8974609/2020\|EPI_ISL_744831\| | ···················· | ···················· | ····················· | ···················· | ···················· | ···················· |
| Z | 919 hCoV-19/Luxembourg/LNS8979333/2020\|EPI_ISL_744832\| | ···················· | ···················· | ····················· | ···················· | ···················· | ···················· |
| Z | 920 hCoV-19/Luxembourg/LNS8999350/2020\|EPI_ISL_744833\| | ············T······· | ···················· | ····················· | ···················· | ···················· | ···················· |
| Z | 921 hCoV-19/Luxembourg/LNS9000833/2020\|EPI_ISL_744834\| | ···················· | G··················· | ····················· | ···················· | ···················· | ···················· |
| Z | 922 hCoV-19/Luxembourg/LNS9007346/2020\|EPI_ISL_744835\| | ···················· | ···················· | ····················· | ···················· | ···················· | ···················· |
| Z | 923 hCoV-19/Luxembourg/LNS9025132/2020\|EPI_ISL_744836\| | ···················· | ···················· | ····················· | ···················· | ···················· | ···················· |
| Z | 924 hCoV-19/Luxembourg/LNS9045971/2020\|EPI_ISL_744837\| | ···················· | ···················· | ····················· | ···················· | ···················· | ···················· |
| Z | 925 hCoV-19/Luxembourg/LNS9049294/2020\|EPI_ISL_744838\| | ···················· | ···················· | ····················· | ···················· | ···················· | ···················· |
| Z | 926 hCoV-19/Luxembourg/LNS9060494/2020\|EPI_ISL_744839\| | ···················· | ···················· | ····················· | ···················· | ···················· | ···················· |
| Z | 927 hCoV-19/Luxembourg/LNS9061386/2020\|EPI_ISL_744840\| | ···················· | ···················· | ····················· | ···················· | ···················· | ···················· |
| Z | 928 hCoV-19/Luxembourg/LNS9063055/2020\|EPI_ISL_744841\| | ···················· | ···················· | ····················· | ···················· | ···················· | ···················· |
| Z | 929 hCoV-19/Luxembourg/LNS9068454/2020\|EPI_ISL_744842\| | ···················· | ···················· | ····················· | ···················· | ···················· | ···················· |
| Z | 930 hCoV-19/Luxembourg/LNS9074543/2020\|EPI_ISL_744843\| | ···················· | ···················· | ····················· | ···················· | ···················· | ···················· |
| Z | 931 hCoV-19/Luxembourg/LNS9081291/2020\|EPI_ISL_744844\| | ···················· | ···················· | ····················· | ···················· | ···················· | ···················· |
| Z | 932 hCoV-19/Luxembourg/LNS9084900/2020\|EPI_ISL_744845\| | ···················· | ···················· | ····················· | ···················· | ···················· | ···················· |
| Z | 933 hCoV-19/Luxembourg/LNS9092417/2020\|EPI_ISL_744847\| | ···················· | G··················· | ····················· | ···················· | ···················· | ···················· |
| Z | 934 hCoV-19/Luxembourg/LNS9095238/2020\|EPI_ISL_744848\| | ···················· | ···················· | ····················· | ···················· | ···················· | ···················· |
| Z | 935 hCoV-19/Luxembourg/LNS9096216/2020\|EPI_ISL_744849\| | ···················· | ···················· | ····················· | ···················· | ···················· | ···················· |
| Z | 936 hCoV-19/Luxembourg/LNS9100060/2020\|EPI_ISL_744852\| | ···················· | ···················· | ····················· | ···················· | ···················· | ···················· |
| Z | 937 hCoV-19/Luxembourg/LNS9101389/2020\|EPI_ISL_744853\| | ···················· | ···················· | ····················· | ···················· | ···················· | ···················· |
| Z | 938 hCoV-19/Luxembourg/LNS9101888/2020\|EPI_ISL_744854\| | ···················· | ···················· | ····················· | ···················· | ···················· | ···················· |
| Z | 939 hCoV-19/Luxembourg/LNS9117076/2020\|EPI_ISL_744855\| | ···················· | ···················· | ····················· | ···················· | ···················· | ···················· |
| Z | 940 hCoV-19/Luxembourg/LNS9202819/2020\|EPI_ISL_744868\| | ···················· | ···················· | ····················· | ···················· | ···················· | ···················· |
| Z | 941 hCoV-19/Luxembourg/LNS9211664/2020\|EPI_ISL_744869\| | ···················· | ···················· | ····················· | ···················· | ···················· | ···················· |
| Z | 942 hCoV-19/Luxembourg/LNS9226115/2020\|EPI_ISL_744871\| | ···················· | ···················· | ····················· | ···················· | ···················· | ···················· |
| Z | 943 hCoV-19/Luxembourg/LNS9231319/2020\|EPI_ISL_744872\| | ···················· | ···················· | ····················· | ···················· | ···················· | ···················· |
| Z | 944 hCoV-19/Luxembourg/LNS9233565/2020\|EPI_ISL_744873\| | ···················· | ···················· | ····················· | ···················· | ···················· | ···················· |
| Z | 945 hCoV-19/Luxembourg/LNS9245793/2020\|EPI_ISL_744874\| | ···················· | ···················· | ····················· | ···················· | ···················· | ···················· |
| Z | 946 hCoV-19/Luxembourg/LNS9255622/2020\|EPI_ISL_744875\| | ···················· | ···················· | ····················· | ···················· | ···················· | ···················· |
| Z | 947 hCoV-19/Luxembourg/LNS9256242/2020\|EPI_ISL_744876\| | ···················· | ···················· | ····················· | ···················· | ······T············· | ···················· |
| Z | 948 hCoV-19/Luxembourg/LNS9257188/2020\|EPI_ISL_744877\| | ···················· | ···················· | ····················· | ···················· | ···················· | ···················· |
| Z | 949 hCoV-19/Luxembourg/LNS9262171/2020\|EPI_ISL_744878\| | ···················· | ···················· | ····················· | ···················· | ···················· | ···················· |
| Z | 950 hCoV-19/Luxembourg/LNS9262856/2020\|EPI_ISL_744879\| | ···················· | ····A··············· | ····················· | ···················· | ···················· | ···················· |
| Z | 951 hCoV-19/Luxembourg/LNS9266699/2020\|EPI_ISL_744880\| | ···················· | ···················· | ····················· | ···················· | ···················· | ···················· |
| Z | 952 hCoV-19/Luxembourg/LNS9304589/2020\|EPI_ISL_744881\| | ···················· | ···················· | ····················· | ···················· | ···················· | ···················· |
| Z | 953 hCoV-19/Luxembourg/LNS9305404/2020\|EPI_ISL_744882\| | ···················· | ···················· | ····················· | ···················· | ···················· | ···················· |
| Z | 954 hCoV-19/Luxembourg/LNS9315713/2020\|EPI_ISL_744883\| | ···················· | ···················· | ····················· | ···················· | ······T············· | ···················· |
| Z | 955 hCoV-19/Luxembourg/LNS9326809/2020\|EPI_ISL_744884\| | ···················· | ···················· | ····················· | ···················· | ···················· | ···················· |
| Z | 956 hCoV-19/Luxembourg/LNS9330908/2020\|EPI_ISL_744885\| | ···················· | ···················· | ····················· | ···················· | ···················· | ···················· |
| Z | 957 hCoV-19/Luxembourg/LNS9338692/2020\|EPI_ISL_744886\| | ···················· | ···················· | ····················· | ···················· | ···················· | ···················· |
| Z | 958 hCoV-19/Luxembourg/LNS9345920/2020\|EPI_ISL_744887\| | ···················· | ···················· | ····················· | ···················· | ···················· | ···················· |
| Z | 959 hCoV-19/Luxembourg/LNS9346393/2020\|EPI_ISL_744888\| | ···················· | ···················· | ····················· | ···················· | ···················· | ···················· |
| Z | 960 hCoV-19/Luxembourg/LNS9348250/2020\|EPI_ISL_744889\| | ···················· | ···················· | ····················· | ···················· | ···················· | ···················· |
| Z | 961 hCoV-19/Luxembourg/LNS9348261/2020\|EPI_ISL_744890\| | ···················· | ···················· | ····················· | ···················· | ···················· | ···················· |
| Z | 962 hCoV-19/Luxembourg/LNS9358147/2020\|EPI_ISL_744891\| | ···················· | ···················· | ····················· | ···················· | ···················· | ···················· |

| | | | | | | | | |
|---|---|---|---|---|---|---|---|---|
| Z | 963 | hCoV-19/Luxembourg/LNS9361630/2020\|EPI_ISL_744892\| | ···················· | ···················· | ····················· | ···················· | ···················· | ···················· |
| Z | 964 | hCoV-19/Luxembourg/LNS9363637/2020\|EPI_ISL_744893\| | ···················· | ···················· | ····················· | ···················· | ···················· | ···················· |
| Z | 965 | hCoV-19/Luxembourg/LNS9375963/2020\|EPI_ISL_744895\| | ···················· | ···················· | ····················· | ···················· | ···················· | ···················· |
| Z | 966 | hCoV-19/Luxembourg/LNS9383265/2020\|EPI_ISL_744896\| | ···················· | ···················· | ····················· | ···················· | ···················· | ···················· |
| Z | 967 | hCoV-19/Luxembourg/LNS9385975/2020\|EPI_ISL_744897\| | ···················· | ···················· | ····················· | ···················· | ···················· | ···················· |
| Z | 968 | hCoV-19/Luxembourg/LNS9386841/2020\|EPI_ISL_744898\| | ···················· | ···················· | ····················· | ···················· | ···················· | ···················· |
| Z | 969 | hCoV-19/Luxembourg/LNS9388233/2020\|EPI_ISL_744899\| | ···················· | ···················· | ····················· | ···················· | ···················· | ···················· |
| Z | 970 | hCoV-19/Luxembourg/LNS9388612/2020\|EPI_ISL_744900\| | ···················· | ···················· | ····················· | ···················· | ···················· | ···················· |
| Z | 971 | hCoV-19/Luxembourg/LNS9388829/2020\|EPI_ISL_744901\| | ···················· | ···················· | ····················· | ···················· | ······T············· | ···················· |
| Z | 972 | hCoV-19/Luxembourg/LNS9394139/2020\|EPI_ISL_744902\| | ···················· | ···················· | ····················· | ···················· | ···················· | ···················· |
| Z | 973 | hCoV-19/Luxembourg/LNS9402077/2020\|EPI_ISL_744904\| | ···················· | ···················· | ····················· | ···················· | ···················· | ···················· |
| Z | 974 | hCoV-19/Luxembourg/LNS9405244/2020\|EPI_ISL_744905\| | ···················· | ···················· | ····················· | ···················· | ···················· | ···················· |
| Z | 975 | hCoV-19/Luxembourg/LNS9407672/2020\|EPI_ISL_744906\| | ···················· | ···················· | ····················· | ···················· | ···················· | ···················· |
| Z | 976 | hCoV-19/Luxembourg/LNS9414120/2020\|EPI_ISL_744907\| | ···················· | ···················· | ····················· | ···················· | ···················· | ···················· |
| Z | 977 | hCoV-19/Luxembourg/LNS9419524/2020\|EPI_ISL_744909\| | ···················· | ···················· | ····················· | ···················· | ···················· | ···················· |
| Z | 978 | hCoV-19/Luxembourg/LNS9425421/2020\|EPI_ISL_744911\| | ···················· | ···················· | ····················· | ···················· | ···················· | ···················· |
| Z | 979 | hCoV-19/Luxembourg/LNS9426550/2020\|EPI_ISL_744912\| | ···················· | ···················· | ····················· | ···················· | ···················· | ···················· |
| Z | 980 | hCoV-19/Luxembourg/LNS9432832/2020\|EPI_ISL_744913\| | ···················· | ···················· | ····················· | ···················· | ···················· | ···················· |
| Z | 981 | hCoV-19/Luxembourg/LNS9434917/2020\|EPI_ISL_744914\| | ···················· | ···················· | ····················· | ···················· | ···················· | ···················· |
| Z | 982 | hCoV-19/Luxembourg/LNS9443496/2020\|EPI_ISL_744915\| | ···················· | ···················· | ····················· | ···················· | ···················· | ···················· |
| Z | 983 | hCoV-19/Luxembourg/LNS9452423/2020\|EPI_ISL_744916\| | ···················· | G··················· | ····················· | ···················· | ···················· | ···················· |
| Z | 984 | hCoV-19/Luxembourg/LNS9454138/2020\|EPI_ISL_744917\| | ···················· | ···················· | ····················· | ···················· | ···················· | ···················· |
| Z | 985 | hCoV-19/Luxembourg/LNS9455907/2020\|EPI_ISL_744918\| | ···················· | ···················· | ····················· | ···················· | ···················· | ···················· |
| Z | 986 | hCoV-19/Luxembourg/LNS9459476/2020\|EPI_ISL_744919\| | ···················· | ···················· | ····················· | ···················· | ···················· | ···················· |
| Z | 987 | hCoV-19/Luxembourg/LNS9462606/2020\|EPI_ISL_744920\| | ···················· | ···················· | ····················· | ···················· | ···················· | ···················· |
| Z | 988 | hCoV-19/Luxembourg/LNS9465071/2020\|EPI_ISL_744921\| | ···················· | ···················· | ····················· | ···················· | ···················· | ···················· |
| Z | 989 | hCoV-19/Luxembourg/LNS9468406/2020\|EPI_ISL_744922\| | ···················· | ···················· | ····················· | ···················· | ···················· | ···················· |
| Z | 990 | hCoV-19/Luxembourg/LNS9475401/2020\|EPI_ISL_744923\| | ···················· | ···················· | ····················· | ···················· | ···················· | ···················· |
| Z | 991 | hCoV-19/Luxembourg/LNS9480033/2020\|EPI_ISL_744924\| | ···················· | ···················· | ····················· | ···················· | ···················· | ···················· |
| Z | 992 | hCoV-19/Luxembourg/LNS9491271/2020\|EPI_ISL_744925\| | ···················· | ···················· | ····················· | ···················· | ···················· | ···················· |
| Z | 993 | hCoV-19/Luxembourg/LNS9502123/2020\|EPI_ISL_744926\| | ···················· | ···················· | ····················· | ···················· | ···················· | ···················· |
| Z | 994 | hCoV-19/Luxembourg/LNS9506014/2020\|EPI_ISL_744927\| | ···················· | ···················· | ····················· | ···················· | ···················· | ···················· |
| Z | 995 | hCoV-19/Luxembourg/LNS9514951/2020\|EPI_ISL_744928\| | ···················· | ···················· | ····················· | ···················· | ···················· | ···················· |
| Z | 996 | hCoV-19/Luxembourg/LNS9518531/2020\|EPI_ISL_744929\| | ···················· | ···················· | ····················· | ···················· | ···················· | ···················· |
| Z | 997 | hCoV-19/Luxembourg/LNS9520185/2020\|EPI_ISL_744930\| | ···················· | ···················· | ····················· | ···················· | ···················· | ···················· |
| Z | 998 | hCoV-19/Luxembourg/LNS9520401/2020\|EPI_ISL_744931\| | ···················· | ···················· | ····················· | ···················· | ···················· | ···················· |
| Z | 999 | hCoV-19/Luxembourg/LNS9525681/2020\|EPI_ISL_744932\| | ···················· | ···················· | ····················· | ···················· | ···················· | ···················· |
| Z | 1000 | hCoV-19/Luxembourg/LNS9528190/2020\|EPI_ISL_744933\| | ···················· | ···················· | ····················· | ···················· | ···················· | ···················· |
| Z | 1001 | hCoV-19/Luxembourg/LNS9530124/2020\|EPI_ISL_744934\| | ···················· | ···················· | ····················· | ···················· | ······T············· | ···················· |
| Z | 1002 | hCoV-19/Luxembourg/LNS9556492/2020\|EPI_ISL_744935\| | ···················· | ···················· | ····················· | ···················· | ···················· | ···················· |
| Z | 1003 | hCoV-19/Luxembourg/LNS9557819/2020\|EPI_ISL_744936\| | ···················· | ·A·················· | ····················· | ···················· | ···················· | ···················· |
| Z | 1004 | hCoV-19/Luxembourg/LNS9559562/2020\|EPI_ISL_744937\| | ···················· | ···················· | ····················· | ···················· | ···················· | ···················· |
| Z | 1005 | hCoV-19/Luxembourg/LNS9561167/2020\|EPI_ISL_744938\| | ···················· | ···················· | ····················· | ···················· | ···················· | ···················· |
| Z | 1006 | hCoV-19/Luxembourg/LNS9561606/2020\|EPI_ISL_744939\| | ···················· | ···················· | ····················· | ···················· | ···················· | ···················· |
| Z | 1007 | hCoV-19/Luxembourg/LNS9561740/2020\|EPI_ISL_744940\| | ···················· | ···················· | ····················· | ···················· | ···················· | ···················· |
| Z | 1008 | hCoV-19/Luxembourg/LNS9564556/2020\|EPI_ISL_744941\| | ···················· | ·A·················· | ····················· | ···················· | ···················· | ···················· |
| Z | 1009 | hCoV-19/Luxembourg/LNS9572611/2020\|EPI_ISL_744942\| | ···················· | ···················· | ····················· | ···················· | ···················· | ···················· |
| Z | 1010 | hCoV-19/Luxembourg/LNS9580764/2020\|EPI_ISL_744943\| | ···················· | ···················· | ····················· | ···················· | ···················· | ···················· |
| Z | 1011 | hCoV-19/Luxembourg/LNS9700021/2020\|EPI_ISL_744970\| | ···················· | ···················· | ····················· | ···················· | ···················· | ···················· |
| Z | 1012 | hCoV-19/Luxembourg/LNS9726231/2020\|EPI_ISL_744972\| | ···················· | ···················· | ····················· | ···················· | ···················· | ···················· |
| Z | 1013 | hCoV-19/Luxembourg/LNS9737544/2020\|EPI_ISL_744975\| | ···················· | ···················· | ····················· | ···················· | ···················· | ···················· |
| Z | 1014 | hCoV-19/Luxembourg/LNS9741818/2020\|EPI_ISL_744976\| | ···················· | ···················· | ····················· | ···················· | ···················· | ···················· |
| Z | 1015 | hCoV-19/Luxembourg/LNS9749175/2020\|EPI_ISL_744977\| | ···················· | ···················· | ····················· | ···················· | ···················· | ···················· |
| Z | 1016 | hCoV-19/Luxembourg/LNS9752865/2020\|EPI_ISL_744978\| | ···················· | ···················· | ····················· | ···················· | ···················· | ···················· |
| Z | 1017 | hCoV-19/Luxembourg/LNS9756090/2020\|EPI_ISL_744979\| | ···················· | ···················· | ····················· | ···················· | ···················· | ···················· |
| Z | 1018 | hCoV-19/Luxembourg/LNS9766242/2020\|EPI_ISL_744980\| | ···················· | ···················· | ····················· | ···················· | ···················· | ···················· |
| Z | 1019 | hCoV-19/Luxembourg/LNS9779064/2020\|EPI_ISL_744981\| | ···················· | ···················· | ····················· | ···················· | ···················· | ···················· |
| Z | 1020 | hCoV-19/Luxembourg/LNS9788943/2020\|EPI_ISL_744982\| | ···················· | ···················· | ····················· | ···················· | ···················· | ···················· |
| Z | 1021 | hCoV-19/Luxembourg/LNS9789059/2020\|EPI_ISL_744983\| | ···················· | ···················· | ····················· | ···················· | ···················· | ···················· |
| Z | 1022 | hCoV-19/Luxembourg/LNS9789433/2020\|EPI_ISL_744984\| | ···················· | ···················· | ····················· | ···················· | ···················· | ···················· |
| Z | 1023 | hCoV-19/Luxembourg/LNS9794728/2020\|EPI_ISL_744985\| | ···················· | ···················· | ····················· | ···················· | ······T············· | ···················· |
| Z | 1024 | hCoV-19/Luxembourg/LNS9795610/2020\|EPI_ISL_744986\| | ···················· | ···················· | ····················· | ···················· | ···················· | ···················· |
| Z | 1025 | hCoV-19/Luxembourg/LNS0000203/2020\|EPI_ISL_739693\| | ···················· | ···················· | ····················· | ···················· | ···················· | ···················· |
| Z | 1026 | hCoV-19/Luxembourg/LNS0003139/2020\|EPI_ISL_739694\| | ···················· | ···················· | ····················· | ···················· | ···················· | ···················· |
| Z | 1027 | hCoV-19/Luxembourg/LNS0005100/2020\|EPI_ISL_739695\| | ···················· | ···················· | ····················· | ···················· | ···················· | ···················· |
| Z | 1028 | hCoV-19/Luxembourg/LNS0010795/2020\|EPI_ISL_739696\| | ···················· | ···················· | ····················· | ···················· | ···················· | ···················· |
| Z | 1029 | hCoV-19/Luxembourg/LNS0022046/2020\|EPI_ISL_739697\| | ···················· | ···················· | ····················· | ···················· | ···················· | ···················· |
| Z | 1030 | hCoV-19/Luxembourg/LNS0025790/2020\|EPI_ISL_739698\| | ···················· | ···················· | ····················· | ···················· | ······T············· | ···················· |
| Z | 1031 | hCoV-19/Luxembourg/LNS0028165/2020\|EPI_ISL_739699\| | ···················· | ···················· | ····················· | ···················· | ···················· | ···················· |
| Z | 1032 | hCoV-19/Canada/BC-BCCDC-7296/2020\|EPI_ISL_740665\|2 | ···················· | ···················· | ····················· | ···················· | ···················· | ···················· |
| Z | 1033 | hCoV-19/Canada/BC-BCCDC-7299/2020\|EPI_ISL_740668\|2 | ···················· | ·A·················· | ····················· | ···················· | ···················· | ···A················ |
| Z | 1034 | hCoV-19/Canada/BC-BCCDC-7300/2020\|EPI_ISL_740669\|2 | ···················· | ·A·················· | ····················· | ···················· | ···················· | ···A················ |
| Z | 1035 | hCoV-19/Canada/BC-BCCDC-7301/2020\|EPI_ISL_740670\|2 | ···················· | ·A·················· | ····················· | ···················· | ···················· | ···A················ |
| Z | 1036 | hCoV-19/Canada/BC-BCCDC-7314/2020\|EPI_ISL_740678\|2 | ···················· | ·A·················· | ····················· | ···················· | ···················· | ···A················ |
| Z | 1037 | hCoV-19/Canada/BC-BCCDC-7316/2020\|EPI_ISL_740680\|2 | ···················· | ·A·················· | ····················· | ···················· | ···················· | ···A················ |
| Z | 1038 | hCoV-19/Canada/BC-BCCDC-7318/2020\|EPI_ISL_740681\|2 | ···················· | ·A·················· | ····················· | ···················· | ···················· | ···A················ |
| Z | 1039 | hCoV-19/Canada/BC-BCCDC-7319/2020\|EPI_ISL_740682\|2 | ···················· | ·A·················· | ····················· | ···················· | ···················· | ···A················ |
| Z | 1040 | hCoV-19/Canada/BC-BCCDC-7320/2020\|EPI_ISL_740683\|2 | ···················· | ·A·················· | ···········T········· | ···················· | ···················· | ···A················ |
| Z | 1041 | hCoV-19/Canada/BC-BCCDC-7321/2020\|EPI_ISL_740684\|2 | ···················· | ·A·················· | ····················· | ···················· | ···················· | ···A················ |
| Z | 1042 | hCoV-19/Canada/BC-BCCDC-7510/2020\|EPI_ISL_740685\|2 | ···················· | ···················· | ····················· | ···················· | ···················· | ···················· |

| | | | | | | | |
|---|---|---|---|---|---|---|---|
| Z | 1043 hCoV-19/Canada/BC-BCCDC-7515/2020\|EPI_ISL_740686\|2 | ···················· | ···················· | ····················· | ···················· | ···················· | ···················· |
| Z | 1044 hCoV-19/Canada/BC-BCCDC-7517/2020\|EPI_ISL_740687\|2 | ···················· | ···················· | ····················· | ···················· | ···················· | ···················· |
| Z | 1045 hCoV-19/Canada/BC-BCCDC-7520/2020\|EPI_ISL_740689\|2 | ···················· | ···················· | ····················· | ···················· | ···················· | ···················· |
| Z | 1046 hCoV-19/Canada/BC-BCCDC-7521/2020\|EPI_ISL_740690\|2 | ···················· | ···················· | ····················· | ···················· | ···················· | ···················· |
| Z | 1047 hCoV-19/Canada/BC-BCCDC-7524/2020\|EPI_ISL_740693\|2 | ···················· | ···················· | ····················· | ···················· | ···················· | ···················· |
| Z | 1048 hCoV-19/Canada/BC-BCCDC-7525/2020\|EPI_ISL_740694\|2 | ···················· | ·············A······ | ····················· | ···················· | ···················· | ···················· |
| Z | 1049 hCoV-19/Canada/BC-BCCDC-7527/2020\|EPI_ISL_740695\|2 | ···················· | ·A·················· | ····················· | ···················· | ···················· | ···A················ |
| Z | 1050 hCoV-19/Canada/BC-BCCDC-7528/2020\|EPI_ISL_740696\|2 | ···················· | ·A·················· | ····················· | ···················· | ···················· | ···A················ |
| Z | 1051 hCoV-19/Canada/BC-BCCDC-7529/2020\|EPI_ISL_740697\|2 | ···················· | ·A·················· | ····················· | ···················· | ···················· | ···A················ |
| Z | 1052 hCoV-19/Canada/BC-BCCDC-7530/2020\|EPI_ISL_740698\|2 | ···················· | ·A·················· | ····················· | ···················· | ···················· | ···A················ |
| Z | 1053 hCoV-19/England/CAMB-A97B8B/2020\|EPI_ISL_741000\|20 | ···················· | ···················· | ····················· | ···················· | ···················· | ···················· |
| Z | 1054 hCoV-19/England/CAMB-A97BB8/2020\|EPI_ISL_741001\|20 | ···················· | ···················· | ····················· | ···················· | ······T············· | ···················· |
| Z | 1055 hCoV-19/England/CAMB-A97D76/2020\|EPI_ISL_741009\|20 | ···················· | ···················· | ····················· | ···················· | ······T············· | ···················· |
| Z | 1057 hCoV-19/England/CAMB-A97EFB/2020\|EPI_ISL_741014\|20 | ···················· | ···················· | ····················· | ···················· | ···················· | ···················· |
| Z | 1058 hCoV-19/England/CAMC-B7982D/2020\|EPI_ISL_741018\|20 | ···················· | ···················· | ····················· | ···················· | ···················· | ···················· |
| Z | 1059 hCoV-19/England/CAMC-B7983C/2020\|EPI_ISL_741019\|20 | ···················· | ···················· | ····················· | ···················· | ···················· | ···················· |
| Z | 1060 hCoV-19/England/CAMC-B7984B/2020\|EPI_ISL_741020\|20 | ···················· | ···················· | ····················· | ···················· | ···················· | ···················· |
| Z | 1061 hCoV-19/England/CAMC-B7985A/2020\|EPI_ISL_741021\|20 | ···················· | ···················· | ····················· | ···················· | ······T············· | ···················· |
| Z | 1062 hCoV-19/England/CAMC-B79896/2020\|EPI_ISL_741024\|20 | ···················· | ···················· | ····················· | ···················· | ···················· | ···················· |
| Z | 1063 hCoV-19/England/CAMC-B798A5/2020\|EPI_ISL_741025\|20 | ···················· | ···················· | ····················· | ···················· | ···················· | ···················· |
| Z | 1064 hCoV-19/England/CAMC-B798B4/2020\|EPI_ISL_741026\|20 | ···················· | ···················· | ····················· | ···················· | ···················· | ···················· |
| Z | 1065 hCoV-19/England/CAMC-B798D2/2020\|EPI_ISL_741027\|20 | ···················· | ···················· | ····················· | ···················· | ···················· | ···················· |
| Z | 1066 hCoV-19/England/CAMC-B798E1/2020\|EPI_ISL_741028\|20 | ···················· | ···················· | ····················· | ···················· | ···················· | ···················· |
| Z | 1067 hCoV-19/England/CAMC-B7991B/2020\|EPI_ISL_741030\|20 | ···················· | ···················· | ····················· | ···················· | ···················· | ···················· |
| Z | 1068 hCoV-19/England/CAMC-B79939/2020\|EPI_ISL_741031\|20 | ···················· | ···················· | ····················· | ···················· | ···················· | ···················· |
| Z | 1069 hCoV-19/England/CAMC-B79A63/2020\|EPI_ISL_741032\|20 | ···················· | ···················· | ····················· | ···················· | ···················· | ···················· |
| Z | 1070 hCoV-19/England/CAMC-B79A72/2020\|EPI_ISL_741033\|20 | ···················· | ···················· | ····················· | ···················· | ···················· | ···················· |
| Z | 1071 hCoV-19/England/CAMC-B79A90/2020\|EPI_ISL_741034\|20 | ···················· | ·A·················· | ····················· | ···················· | ···················· | ···················· |
| Z | 1072 hCoV-19/England/CAMC-B79AAF/2020\|EPI_ISL_741035\|20 | ···················· | ···················· | ····················· | ···················· | ···················· | ···················· |
| Z | 1073 hCoV-19/England/CAMC-B79ABE/2020\|EPI_ISL_741036\|20 | ········T··········· | ···················· | ····················· | ···················· | ···················· | ···················· |
| Z | 1074 hCoV-19/England/CAMC-B79BF7/2020\|EPI_ISL_741038\|20 | ···················· | ···················· | ····················· | ···················· | ···················· | ···················· |
| Z | 1075 hCoV-19/England/CAMC-B79C12/2020\|EPI_ISL_741040\|20 | ···················· | ···················· | ····················· | ···················· | ···················· | ···················· |
| Z | 1076 hCoV-19/England/CAMC-B79C21/2020\|EPI_ISL_741041\|20 | ···················· | ···················· | ····················· | ···················· | ···················· | ···················· |
| Z | 1077 hCoV-19/England/CAMC-B79C4F/2020\|EPI_ISL_741043\|20 | ···················· | ···················· | ····················· | ···················· | ···················· | ···················· |
| Z | 1078 hCoV-19/England/CAMC-B79C9A/2020\|EPI_ISL_741047\|20 | ···················· | ···················· | ····················· | ···················· | ······T············· | ···················· |
| Z | 1079 hCoV-19/England/CAMC-B79E2B/2020\|EPI_ISL_741051\|20 | ···················· | ···················· | ····················· | ···················· | ···················· | ···················· |
| Z | 1080 hCoV-19/England/CAMC-B79E3A/2020\|EPI_ISL_741052\|20 | ···················· | ···················· | ····················· | ···················· | ···················· | ···················· |
| Z | 1081 hCoV-19/England/CAMC-B79E58/2020\|EPI_ISL_741053\|20 | ···················· | ···················· | ····················· | ···················· | ···················· | ···················· |
| Z | 1082 hCoV-19/England/CAMC-B79E67/2020\|EPI_ISL_741054\|20 | ···················· | ···················· | ····················· | ···················· | ···················· | ···················· |
| Z | 1083 hCoV-19/England/CAMC-B79E76/2020\|EPI_ISL_741055\|20 | ···················· | ···················· | ····················· | ···················· | ···················· | ···················· |
| Z | 1084 hCoV-19/England/CAMC-B79E85/2020\|EPI_ISL_741056\|20 | ···················· | ·A·················· | ····················· | ···················· | ···················· | ···A················ |
| Z | 1085 hCoV-19/England/CAMC-B79E94/2020\|EPI_ISL_741057\|20 | ···················· | ···················· | ····················· | ···················· | ···················· | ···················· |
| Z | 1086 hCoV-19/England/CAMC-B79EB2/2020\|EPI_ISL_741059\|20 | ···················· | ···················· | ····················· | ···················· | ···················· | ···················· |
| Z | 1087 hCoV-19/England/CAMC-B79EC1/2020\|EPI_ISL_741060\|20 | ···················· | ···················· | ····················· | ···················· | ···················· | ···················· |
| Z | 1088 hCoV-19/England/CAMC-B79F0A/2020\|EPI_ISL_741062\|20 | ···················· | ···················· | ····················· | ···················· | ···················· | ···················· |
| Z | 1089 hCoV-19/England/CAMC-B79F19/2020\|EPI_ISL_741063\|20 | ···················· | ···················· | ····················· | ···················· | ···················· | ···················· |
| Z | 1090 hCoV-19/England/CAMC-B79F28/2020\|EPI_ISL_741064\|20 | ···················· | ···················· | ····················· | ···················· | ···················· | ···················· |
| Z | 1091 hCoV-19/England/CAMC-B79F37/2020\|EPI_ISL_741065\|20 | ···················· | ·A·················· | ····················· | ···················· | ···················· | ···A················ |
| Z | 1092 hCoV-19/England/CAMC-B79F46/2020\|EPI_ISL_741066\|20 | ···················· | ···················· | ····················· | ···················· | ···················· | ···················· |
| Z | 1093 hCoV-19/England/CAMC-B79F73/2020\|EPI_ISL_741068\|20 | ···················· | ···················· | ····················· | ···················· | ···················· | ···················· |
| Z | 1094 hCoV-19/England/CAMC-B79F82/2020\|EPI_ISL_741069\|20 | ···················· | ···················· | ····················· | ···················· | ···················· | ···················· |
| Z | 1095 hCoV-19/England/CAMC-B79F91/2020\|EPI_ISL_741070\|20 | ···················· | ···················· | ····················· | ···················· | ···················· | ···················· |
| Z | 1096 hCoV-19/England/CAMC-B79FCE/2020\|EPI_ISL_741073\|20 | ···················· | ···················· | ····················· | ···················· | ···················· | ···················· |
| Z | 1097 hCoV-19/England/CAMC-B79FEC/2020\|EPI_ISL_741074\|20 | ···················· | ···················· | ····················· | ···················· | ···················· | ···················· |
| Z | 1098 hCoV-19/England/CAMC-B7A006/2020\|EPI_ISL_741075\|20 | ···················· | ···················· | ····················· | ···················· | ···················· | ···················· |
| Z | 1099 hCoV-19/England/CAMC-B7A024/2020\|EPI_ISL_741076\|20 | ···················· | ···················· | ····················· | ···················· | ······T············· | ···················· |
| Z | 1100 hCoV-19/England/CAMC-B7A033/2020\|EPI_ISL_741077\|20 | ···················· | ···················· | ····················· | ···················· | ···················· | ···················· |
| Z | 1101 hCoV-19/England/CAMC-B7A042/2020\|EPI_ISL_741078\|20 | ···················· | ···················· | ····················· | ···················· | ···················· | ···················· |
| Z | 1102 hCoV-19/England/CAMC-B7A051/2020\|EPI_ISL_741079\|20 | ···················· | ···················· | ····················· | ···················· | ···················· | ···················· |
| Z | 1103 hCoV-19/England/CAMC-B7A0CA/2020\|EPI_ISL_741080\|20 | ···················· | ···················· | ····················· | ···················· | ···················· | ···················· |
| Z | 1104 hCoV-19/England/CAMC-B7A0D9/2020\|EPI_ISL_741081\|20 | ···················· | ···················· | ····················· | ···················· | ···················· | ···················· |
| Z | 1105 hCoV-19/England/CAMC-B7A0E8/2020\|EPI_ISL_741082\|20 | ···················· | ···················· | ····················· | ···················· | ···················· | ···················· |
| Z | 1106 hCoV-19/England/CAMC-B7A121/2020\|EPI_ISL_741084\|20 | ···········T········ | ···················· | ····················· | ···················· | ···················· | ···················· |
| Z | 1107 hCoV-19/England/CAMC-B7A16D/2020\|EPI_ISL_741085\|20 | ···················· | ···················· | ····················· | ···················· | ···················· | ···················· |
| Z | 1108 hCoV-19/England/CAMC-B7A17C/2020\|EPI_ISL_741086\|20 | ···················· | ···················· | ····················· | ···················· | ···················· | ···················· |
| Z | 1109 hCoV-19/England/CAMC-B7A1F4/2020\|EPI_ISL_741087\|20 | ···················· | ···················· | ····················· | ···················· | ···················· | ···················· |
| Z | 1110 hCoV-19/England/CAMC-B7A200/2020\|EPI_ISL_741088\|20 | ···················· | ···················· | ····················· | ···················· | ···················· | ···················· |
| Z | 1111 hCoV-19/England/CAMC-B7A21F/2020\|EPI_ISL_741089\|20 | ···················· | ···················· | ····················· | ···················· | ······T············· | ···················· |
| Z | 1112 hCoV-19/England/CAMC-B7A22E/2020\|EPI_ISL_741090\|20 | ···················· | ···················· | ····················· | ···················· | ···················· | ···················· |
| Z | 1113 hCoV-19/England/CAMC-B7A23D/2020\|EPI_ISL_741091\|20 | ···················· | ···················· | ····················· | ···················· | ······T············· | ···················· |
| Z | 1114 hCoV-19/England/CAMC-B7A24C/2020\|EPI_ISL_741092\|20 | ···················· | ···················· | ····················· | ···················· | ···················· | ···················· |
| Z | 1115 hCoV-19/England/CAMC-B7A2A6/2020\|EPI_ISL_741094\|20 | ···················· | ···················· | ····················· | ···················· | ···················· | ···················· |
| Z | 1116 hCoV-19/England/CAMC-B7A2E2/2020\|EPI_ISL_741096\|20 | ···················· | ···················· | ····················· | ···················· | ···················· | ···················· |
| Z | 1117 hCoV-19/England/CAMC-B7A30D/2020\|EPI_ISL_741097\|20 | ···················· | ···················· | ····················· | ···················· | ···················· | ···················· |
| Z | 1118 hCoV-19/England/CAMC-B7A31C/2020\|EPI_ISL_741098\|20 | ···················· | ···················· | ····················· | ···················· | ···················· | ···················· |
| Z | 1119 hCoV-19/England/CAMC-B7A32B/2020\|EPI_ISL_741099\|20 | ···················· | ···················· | ····················· | ···················· | ···················· | ···················· |
| Z | 1120 hCoV-19/England/SHEF-10CCE21/2020\|EPI_ISL_741996\|2 | ···················· | ···················· | ····················· | ···················· | ···················· | ···················· |
| Z | 1121 hCoV-19/Wales/PHWC-49B130/2020\|EPI_ISL_742300\|2020 | ···················· | ···················· | ····················· | ···················· | ···················· | ···················· |
| Z | 1122 hCoV-19/Wales/PHWC-49B14F/2020\|EPI_ISL_742301\|2020 | ···················· | ···················· | ····················· | ···················· | ···················· | ···················· |
| Z | 1123 hCoV-19/Wales/PHWC-49B17C/2020\|EPI_ISL_742303\|2020 | ···················· | ···················· | ····················· | ···················· | ···················· | ···················· |

| | | | | | | | | |
|---|---|---|---|---|---|---|---|---|
| Z | 1124 | hCoV-19/Wales/PHWC-49B18B/2020\|EPI_ISL_742304\|2020 | ···················· | ···················· | ····················· | ···················· | ···················· | ···················· |
| Z | 1125 | hCoV-19/Wales/PHWC-49B19A/2020\|EPI_ISL_742305\|2020 | ···················· | ···················· | ····················· | ···················· | ···················· | ···R················ |
| Z | 1126 | hCoV-19/Wales/PHWC-49B1B8/2020\|EPI_ISL_742306\|2020 | ···················· | ···················· | ····················· | ···················· | ···················· | ···················· |
| Z | 1127 | hCoV-19/Wales/PHWC-49B1C7/2020\|EPI_ISL_742307\|2020 | ···················· | ···················· | ····················· | ···················· | ···················· | ···················· |
| Z | 1128 | hCoV-19/Wales/PHWC-49B1E5/2020\|EPI_ISL_742308\|2020 | ···················· | ···················· | ····················· | ···················· | ···················· | ···················· |
| Z | 1129 | hCoV-19/Wales/PHWC-49B26A/2020\|EPI_ISL_742311\|2020 | ···················· | ···················· | ····················· | ···················· | ···················· | ···················· |
| Z | 1130 | hCoV-19/Wales/PHWC-49B288/2020\|EPI_ISL_742313\|2020 | ···················· | ···················· | ····················· | ···················· | ···················· | ···················· |
| Z | 1131 | hCoV-19/Wales/PHWC-49B297/2020\|EPI_ISL_742314\|2020 | ···················· | ···················· | ····················· | ···················· | ···················· | ···················· |
| Z | 1132 | hCoV-19/Wales/PHWC-49B394/2020\|EPI_ISL_742326\|2020 | ···················· | ···················· | ····················· | ···················· | ···················· | ···················· |
| Z | 1133 | hCoV-19/Wales/PHWC-49B3A3/2020\|EPI_ISL_742327\|2020 | ···················· | ···················· | ····················· | ···················· | ···················· | ···················· |
| Z | 1134 | hCoV-19/Wales/PHWC-49B3B2/2020\|EPI_ISL_742328\|2020 | ···················· | ···················· | ····················· | ···················· | ···················· | ···················· |
| Z | 1135 | hCoV-19/Wales/PHWC-49B3D0/2020\|EPI_ISL_742330\|2020 | ···················· | ···················· | ····················· | ···················· | ···················· | ···················· |
| Z | 1136 | hCoV-19/Wales/PHWC-49B3EF/2020\|EPI_ISL_742331\|2020 | ···················· | ···················· | ····················· | ···················· | ···················· | ···················· |
| Z | 1137 | hCoV-19/Wales/PHWC-49B3FE/2020\|EPI_ISL_742332\|2020 | ···················· | ···················· | ····················· | ···················· | ···················· | ···················· |
| Z | 1138 | hCoV-19/Wales/PHWC-49B40A/2020\|EPI_ISL_742333\|2020 | ···················· | ···················· | ····················· | ···················· | ···················· | ···················· |
| Z | 1139 | hCoV-19/Wales/PHWC-49B419/2020\|EPI_ISL_742334\|2020 | ···················· | ···················· | ····················· | ···················· | ···················· | ···················· |
| Z | 1140 | hCoV-19/Wales/PHWC-49B428/2020\|EPI_ISL_742335\|2020 | ···················· | ···················· | ····················· | ···················· | ···················· | ···················· |
| Z | 1141 | hCoV-19/Wales/PHWC-49B437/2020\|EPI_ISL_742336\|2020 | ···················· | ···················· | ····················· | ···················· | ···················· | ···················· |
| Z | 1142 | hCoV-19/Wales/PHWC-49B446/2020\|EPI_ISL_742337\|2020 | ···················· | ···················· | ····················· | ···················· | ···················· | ···················· |
| Z | 1143 | hCoV-19/Wales/PHWC-49B464/2020\|EPI_ISL_742339\|2020 | ···················· | ···················· | ····················· | ···················· | ···················· | ···················· |
| Z | 1144 | hCoV-19/Wales/PHWC-49B473/2020\|EPI_ISL_742340\|2020 | ···················· | ···················· | ····················· | ···················· | ···················· | ···················· |
| Z | 1145 | hCoV-19/Wales/PHWC-49B482/2020\|EPI_ISL_742341\|2020 | ···················· | ···················· | ····················· | ···················· | ···················· | ···················· |
| Z | 1146 | hCoV-19/Wales/PHWC-49B4A0/2020\|EPI_ISL_742342\|2020 | ···················· | ···················· | ····················· | ···················· | ···················· | ···················· |
| Z | 1147 | hCoV-19/Wales/PHWC-49B4CE/2020\|EPI_ISL_742344\|2020 | ···················· | ···················· | ····················· | ···················· | ···················· | ···················· |
| Z | 1148 | hCoV-19/Wales/PHWC-49B4DD/2020\|EPI_ISL_742345\|2020 | ···················· | ···················· | ····················· | ···················· | ···················· | ···················· |
| Z | 1149 | hCoV-19/Wales/PHWC-49B4EC/2020\|EPI_ISL_742346\|2020 | ···················· | ···················· | ····················· | ···················· | ···················· | ···················· |
| Z | 1150 | hCoV-19/Wales/PHWC-49B4FB/2020\|EPI_ISL_742347\|2020 | ···················· | ···················· | ····················· | ···················· | ···················· | ···················· |
| Z | 1151 | hCoV-19/Wales/PHWC-49B507/2020\|EPI_ISL_742348\|2020 | ···················· | ···················· | ····················· | ···················· | ···················· | ···················· |
| Z | 1152 | hCoV-19/Wales/PHWC-49B525/2020\|EPI_ISL_742349\|2020 | ···················· | ···················· | ····················· | ···················· | ···················· | ···················· |
| Z | 1153 | hCoV-19/Wales/PHWC-49B534/2020\|EPI_ISL_742350\|2020 | ···················· | ···················· | ····················· | ···················· | ···················· | ···················· |
| Z | 1154 | hCoV-19/Wales/PHWC-49B552/2020\|EPI_ISL_742351\|2020 | ···················· | ···················· | ····················· | ···················· | ···················· | ···················· |
| Z | 1159 | hCoV-19/Wales/PHWC-49B5DA/2020\|EPI_ISL_742358\|2020 | ···················· | ···················· | ····················· | ···················· | ···················· | ···················· |
| Z | 1165 | hCoV-19/Wales/PHWC-49B69B/2020\|EPI_ISL_742367\|2020 | ···················· | ···················· | ····················· | ···················· | ···················· | ···················· |
| Z | 1167 | hCoV-19/Wales/PHWC-49B6B9/2020\|EPI_ISL_742369\|2020 | ···················· | ···················· | ····················· | ···················· | ···················· | ···················· |
| Z | 1170 | hCoV-19/Wales/PHWC-49B6E6/2020\|EPI_ISL_742372\|2020 | ···················· | ···················· | ····················· | ···················· | ···················· | ···················· |
| Z | 1171 | hCoV-19/Wales/PHWC-49B6F5/2020\|EPI_ISL_742373\|2020 | ···················· | ···················· | ····················· | ···················· | ···················· | ···················· |
| Z | 1173 | hCoV-19/Wales/PHWC-49B72F/2020\|EPI_ISL_742376\|2020 | ···················· | ···················· | ····················· | ···················· | ···················· | ···················· |
| Z | 1175 | hCoV-19/Wales/PHWC-49B76B/2020\|EPI_ISL_742380\|2020 | ···················· | ···················· | ····················· | ···················· | ···················· | ···················· |
| Z | 1182 | hCoV-19/Wales/PHWC-49B81D/2020\|EPI_ISL_742390\|2020 | ···················· | ···················· | ····················· | ···················· | ···················· | ···················· |
| Z | 1194 | hCoV-19/Wales/PHWC-4A1331/2020\|EPI_ISL_743657\|2020 | ···················· | ···················· | ····················· | ···················· | ···················· | ···················· |
| Z | 1195 | hCoV-19/Wales/PHWC-4A1340/2020\|EPI_ISL_743658\|2020 | ···················· | ···················· | ····················· | ···················· | ···················· | ···················· |
| Z | 1197 | hCoV-19/Wales/PHWC-4A136E/2020\|EPI_ISL_743660\|2020 | ···················· | ···················· | ····················· | ···················· | ···················· | ···················· |
| Z | 1198 | hCoV-19/Wales/PHWC-4A137D/2020\|EPI_ISL_743661\|2020 | ···················· | ···················· | ····················· | ···················· | ···················· | ···················· |
| Z | 1200 | hCoV-19/Wales/PHWC-4A13B9/2020\|EPI_ISL_743664\|2020 | ···················· | ···················· | ····················· | ···················· | ···················· | ···················· |
| Z | 1201 | hCoV-19/Wales/PHWC-4A13C8/2020\|EPI_ISL_743665\|2020 | ···················· | ···················· | ····················· | ···················· | ···················· | ···················· |
| Z | 1202 | hCoV-19/Wales/PHWC-4A13E6/2020\|EPI_ISL_743667\|2020 | ···················· | ···················· | ····················· | ···················· | ···················· | ···················· |
| Z | 1203 | hCoV-19/Wales/PHWC-4A13F5/2020\|EPI_ISL_743668\|2020 | ···················· | ···················· | ····················· | ···················· | ···················· | ···················· |
| Z | 1204 | hCoV-19/Wales/PHWC-4A1410/2020\|EPI_ISL_743670\|2020 | ···················· | ···················· | ····················· | ···················· | ···················· | ···················· |
| Z | 1205 | hCoV-19/Wales/PHWC-4A142F/2020\|EPI_ISL_743671\|2020 | ···················· | ···················· | ····················· | ···················· | ···················· | ···················· |
| Z | 1206 | hCoV-19/Wales/PHWC-4A143E/2020\|EPI_ISL_743672\|2020 | ···················· | ···················· | ····················· | ···················· | ···················· | ···················· |
| Z | 1208 | hCoV-19/Wales/PHWC-4A147A/2020\|EPI_ISL_743676\|2020 | ···················· | ···················· | ····················· | ···················· | ···················· | ···················· |
| Z | 1209 | hCoV-19/Wales/PHWC-4A1489/2020\|EPI_ISL_743677\|2020 | ···················· | ···················· | ····················· | ···················· | ···················· | ···················· |
| Z | 1210 | hCoV-19/Wales/PHWC-4A1498/2020\|EPI_ISL_743678\|2020 | ···················· | ···················· | ····················· | ···················· | ···················· | ···················· |
| Z | 1212 | hCoV-19/Wales/PHWC-4A14B6/2020\|EPI_ISL_743680\|2020 | ···················· | ···················· | ····················· | ···················· | ···················· | ···················· |
| Z | 1213 | hCoV-19/Wales/PHWC-4A14C5/2020\|EPI_ISL_743681\|2020 | ···················· | ···················· | ····················· | ···················· | ···················· | ···················· |
| Z | 1214 | hCoV-19/Wales/PHWC-4A14E3/2020\|EPI_ISL_743682\|2020 | ···················· | ···················· | ····················· | ···················· | ···················· | ···················· |
| Z | 1215 | hCoV-19/Wales/PHWC-4A14F2/2020\|EPI_ISL_743683\|2020 | ···················· | ···················· | ····················· | ···················· | ···················· | ···················· |
| Z | 1217 | hCoV-19/Wales/PHWC-4A151D/2020\|EPI_ISL_743685\|2020 | ···················· | ···················· | ····················· | ···················· | ···················· | ···················· |
| Z | 1219 | hCoV-19/Wales/PHWC-4A153B/2020\|EPI_ISL_743687\|2020 | ···················· | ···················· | ····················· | ···················· | ···················· | ···················· |
| Z | 1220 | hCoV-19/Wales/PHWC-4A154A/2020\|EPI_ISL_743688\|2020 | ···················· | ···················· | ····················· | ···················· | ···················· | ···················· |
| Z | 1222 | hCoV-19/Wales/PHWC-4A1568/2020\|EPI_ISL_743690\|2020 | ···················· | ···················· | ····················· | ···················· | ···················· | ···················· |
| Z | 1223 | hCoV-19/Wales/PHWC-4A1577/2020\|EPI_ISL_743691\|2020 | ···················· | ···················· | ····················· | ···················· | ···················· | ···················· |
| Z | 1224 | hCoV-19/Wales/PHWC-4A1586/2020\|EPI_ISL_743692\|2020 | ···················· | ···················· | ····················· | ···················· | ···················· | ···················· |
| Z | 1226 | hCoV-19/Wales/PHWC-4A15A4/2020\|EPI_ISL_743694\|2020 | ···················· | ···················· | ····················· | ···················· | ···················· | ···················· |
| Z | 1227 | hCoV-19/Wales/PHWC-4A15B3/2020\|EPI_ISL_743695\|2020 | ···················· | ···················· | ····················· | ···················· | ···················· | ···················· |
| Z | 1228 | hCoV-19/Wales/PHWC-4A15C2/2020\|EPI_ISL_743696\|2020 | ···················· | ···················· | ····················· | ···················· | ···················· | ···················· |
| Z | 1230 | hCoV-19/Wales/PHWC-4A15E0/2020\|EPI_ISL_743698\|2020 | ···················· | ···················· | ····················· | ···················· | ···················· | ···················· |
| Z | 1231 | hCoV-19/Luxembourg/LNS9588266/2020\|EPI_ISL_744944\| | ···················· | ····A··············· | ····················· | ···················· | ···················· | ···················· |
| Z | 1232 | hCoV-19/Luxembourg/LNS9594129/2020\|EPI_ISL_744945\| | ···················· | ···················· | ····················· | ···················· | ···················· | ···················· |
| Z | 1233 | hCoV-19/Luxembourg/LNS9594490/2020\|EPI_ISL_744946\| | ···················· | ···················· | ···········T········· | ···················· | ···················· | ···················· |
| Z | 1234 | hCoV-19/Luxembourg/LNS9600094/2020\|EPI_ISL_744947\| | ···················· | ···················· | ····················· | ···················· | ···················· | ···················· |
| Z | 1235 | hCoV-19/Luxembourg/LNS9602424/2020\|EPI_ISL_744948\| | ···················· | ···················· | ····················· | ···················· | ···················· | ···················· |
| Z | 1236 | hCoV-19/Luxembourg/LNS9610397/2020\|EPI_ISL_744950\| | ···················· | ···················· | ····················· | ···················· | ···················· | ···················· |
| Z | 1237 | hCoV-19/Luxembourg/LNS9610915/2020\|EPI_ISL_744951\| | ···················· | ···················· | ····················· | ···················· | ···················· | ···················· |
| Z | 1238 | hCoV-19/Luxembourg/LNS9623706/2020\|EPI_ISL_744952\| | ···················· | ···················· | ····················· | ···················· | ···················· | ···················· |
| Z | 1239 | hCoV-19/Luxembourg/LNS9628686/2020\|EPI_ISL_744953\| | ···················· | ···················· | ····················· | ···················· | ···················· | ···················· |
| Z | 1240 | hCoV-19/Luxembourg/LNS9629548/2020\|EPI_ISL_744954\| | ···················· | ···················· | ····················· | ···················· | ···················· | ···················· |
| Z | 1241 | hCoV-19/Luxembourg/LNS9629893/2020\|EPI_ISL_744955\| | ···················· | ···················· | ····················· | ···················· | ···················· | ···················· |
| Z | 1242 | hCoV-19/Luxembourg/LNS9632945/2020\|EPI_ISL_744956\| | ···················· | ···················· | ····················· | ···················· | ···················· | ···················· |
| Z | 1243 | hCoV-19/Luxembourg/LNS9643334/2020\|EPI_ISL_744957\| | ···················· | ···················· | ····················· | ···················· | ···················· | ···················· |

| | | | | | | | | |
|---|---|---|---|---|---|---|---|---|
| Z | 1244 | hCoV-19/Luxembourg/LNS9651738/2020\|EPI_ISL_744959\| | ···················· | ···················· | ····················· | ···················· | ···················· | ···················· |
| Z | 1245 | hCoV-19/Luxembourg/LNS9657884/2020\|EPI_ISL_744960\| | ···················· | ···················· | ····················· | ···················· | ···················· | ···················· |
| Z | 1246 | hCoV-19/Luxembourg/LNS9660366/2020\|EPI_ISL_744961\| | ···················· | ···················· | ····················· | ···················· | ···················· | ···················· |
| Z | 1247 | hCoV-19/Luxembourg/LNS9661564/2020\|EPI_ISL_744962\| | ···················· | ···················· | ····················· | ···················· | ···················· | ···················· |
| Z | 1248 | hCoV-19/Luxembourg/LNS9663338/2020\|EPI_ISL_744963\| | ···················· | ···················· | ····················· | ···················· | ···················· | ···················· |
| Z | 1249 | hCoV-19/Luxembourg/LNS9668214/2020\|EPI_ISL_744964\| | ···················· | ···················· | ····················· | ···················· | ···················· | ···················· |
| Z | 1250 | hCoV-19/Luxembourg/LNS9680191/2020\|EPI_ISL_744965\| | ···················· | ···················· | ····················· | ···················· | ···················· | ···················· |
| Z | 1251 | hCoV-19/Luxembourg/LNS9690989/2020\|EPI_ISL_744966\| | ···················· | ···················· | ····················· | ···················· | ···················· | ···················· |
| Z | 1252 | hCoV-19/Luxembourg/LNS9691282/2020\|EPI_ISL_744967\| | ···················· | ···················· | ····················· | ···················· | ···················· | ···················· |
| Z | 1253 | hCoV-19/Luxembourg/LNS9698899/2020\|EPI_ISL_744968\| | ···················· | ···················· | ····················· | ···················· | ···················· | ···················· |
| Z | 1254 | hCoV-19/Luxembourg/LNS9699068/2020\|EPI_ISL_744969\| | ···················· | ···················· | ····················· | ···················· | ···················· | ···················· |
| Z | 1255 | hCoV-19/Luxembourg/LNS9726749/2020\|EPI_ISL_744973\| | ···················· | ···················· | ····················· | ···················· | ···················· | ···················· |
| Z | 1256 | hCoV-19/Luxembourg/LNS9732596/2020\|EPI_ISL_744974\| | ···················· | ···················· | ····················· | ···················· | ···················· | ···················· |
| Z | 1257 | hCoV-19/Luxembourg/LNS9803660/2020\|EPI_ISL_744988\| | ···················· | ···················· | ····················· | ···················· | ···················· | ···················· |
| Z | 1258 | hCoV-19/Luxembourg/LNS9804369/2020\|EPI_ISL_744989\| | ···················· | ···················· | ····················· | ···················· | ···················· | ···················· |
| Z | 1259 | hCoV-19/Luxembourg/LNS9804517/2020\|EPI_ISL_744990\| | ···················· | ···················· | ····················· | ···················· | ···················· | ···················· |
| Z | 1260 | hCoV-19/Luxembourg/LNS9805873/2020\|EPI_ISL_744991\| | ···················· | ···················· | ····················· | ···················· | ···················· | ···················· |
| Z | 1261 | hCoV-19/Luxembourg/LNS9806033/2020\|EPI_ISL_744992\| | ···················· | ···················· | ····················· | ···················· | ···················· | ···················· |
| Z | 1262 | hCoV-19/Luxembourg/LNS9819784/2020\|EPI_ISL_744994\| | ···················· | ···················· | ····················· | ···················· | ···················· | ···················· |
| Z | 1263 | hCoV-19/Luxembourg/LNS9828039/2020\|EPI_ISL_744995\| | ···················· | ···················· | ····················· | ···················· | ···················· | ···················· |
| Z | 1264 | hCoV-19/Luxembourg/LNS9837378/2020\|EPI_ISL_744996\| | ···················· | ···················· | ····················· | ···················· | ···················· | ···················· |
| Z | 1265 | hCoV-19/Luxembourg/LNS9838378/2020\|EPI_ISL_744997\| | ···················· | ···················· | ····················· | ···················· | ···················· | ···················· |
| Z | 1266 | hCoV-19/Luxembourg/LNS9863463/2020\|EPI_ISL_744999\| | ···················· | ···················· | ····················· | ···················· | ···················· | ···················· |
| Z | 1267 | hCoV-19/Chile/RM-83613/2020\|EPI_ISL_746800\|2020-05 | ···················· | ···················· | ····················· | ···················· | ···················· | ···················· |
| Z | 1268 | hCoV-19/Chile/RM-84213/2020\|EPI_ISL_746801\|2020-05 | ···················· | ···················· | ····················· | ···················· | ···················· | ···················· |
| Z | 1269 | hCoV-19/Chile/RM-84393/2020\|EPI_ISL_746802\|2020-05 | ···················· | ···················· | ····················· | ···················· | ···················· | ···················· |
| Z | 1270 | hCoV-19/Chile/RM-88162-TC1/2020\|EPI_ISL_746825\|202 | ···················· | ···················· | ····················· | ···················· | ···················· | ···················· |
| Z | 1272 | hCoV-19/Italy/LAZ-INMI-120/2020\|EPI_ISL_746827\|202 | ···················· | ···················· | ····················· | ···················· | ···················· | ···················· |
| Z | 1273 | hCoV-19/Italy/LAZ-INMI-121/2020\|EPI_ISL_746828\|202 | ···················· | ···················· | ····················· | ···················· | ···················· | ···················· |
| Z | 1275 | hCoV-19/Italy/LAZ-INMI-123/2020\|EPI_ISL_746830\|202 | ···················· | ···················· | ····················· | ···················· | ···················· | ···················· |
| Z | 1276 | hCoV-19/Italy/LAZ-INMI-124/2020\|EPI_ISL_746831\|202 | ···················· | ·A·················· | ····················· | ···················· | ···················· | ···················· |
| Z | 1277 | hCoV-19/Italy/LAZ-INMI-125/2020\|EPI_ISL_746832\|202 | ···················· | ···················· | ····················· | ···················· | ······N············· | ···················· |
| Z | 1278 | hCoV-19/Italy/LAZ-INMI-126/2020\|EPI_ISL_746833\|202 | ···················· | ···················· | ····················· | ···················· | ······N············· | ···················· |
| Z | 1279 | hCoV-19/Italy/LAZ-INMI-127/2020\|EPI_ISL_746834\|202 | ···················· | ···················· | ····················· | ···················· | ···················· | ···················· |
| Z | 1280 | hCoV-19/Italy/LAZ-INMI-128/2020\|EPI_ISL_746835\|202 | ···················· | ···················· | ····················· | ···················· | ······N············· | ···················· |
| Z | 1281 | hCoV-19/Luxembourg/LNS0042548/2020\|EPI_ISL_739700\| | ···················· | ···················· | ····················· | ···················· | ···················· | ···················· |
| Z | 1282 | hCoV-19/Luxembourg/LNS0042857/2020\|EPI_ISL_739701\| | ···················· | ···················· | ····················· | ···················· | ···················· | ···················· |
| Z | 1283 | hCoV-19/Luxembourg/LNS0043293/2020\|EPI_ISL_739702\| | ············T······· | ···················· | ····················· | ···················· | ···················· | ···················· |
| Z | 1284 | hCoV-19/Luxembourg/LNS0059319/2020\|EPI_ISL_739703\| | ···················· | ···················· | ····················· | ···················· | ···················· | ···················· |
| Z | 1285 | hCoV-19/Luxembourg/LNS0061167/2020\|EPI_ISL_739704\| | ···················· | ···················· | ····················· | ···················· | ···················· | ···················· |
| Z | 1286 | hCoV-19/Luxembourg/LNS0067083/2020\|EPI_ISL_739705\| | ···················· | ···················· | ····················· | ···················· | ···················· | ···················· |
| Z | 1287 | hCoV-19/Luxembourg/LNS0070392/2020\|EPI_ISL_739706\| | ···················· | ···················· | ····················· | ···················· | ···················· | ···················· |
| Z | 1288 | hCoV-19/Luxembourg/LNS0074724/2020\|EPI_ISL_739707\| | ···················· | ···················· | ····················· | ···················· | ···················· | ···················· |
| Z | 1289 | hCoV-19/Luxembourg/LNS0084824/2020\|EPI_ISL_739708\| | ···················· | ···················· | ····················· | ···················· | ···················· | ···················· |
| Z | 1290 | hCoV-19/Luxembourg/LNS0087318/2020\|EPI_ISL_739709\| | ···················· | ···················· | ····················· | ···················· | ···················· | ···················· |
| Z | 1291 | hCoV-19/Luxembourg/LNS0100665/2020\|EPI_ISL_739710\| | ···················· | ···················· | ····················· | ···················· | ···················· | ···················· |
| Z | 1292 | hCoV-19/Luxembourg/LNS0102726/2020\|EPI_ISL_739711\| | ···················· | ···················· | ····················· | ···················· | ···················· | ···················· |
| Z | 1293 | hCoV-19/Luxembourg/LNS0104363/2020\|EPI_ISL_739712\| | ···················· | ···················· | ····················· | ···················· | ···················· | ···················· |
| Z | 1294 | hCoV-19/Luxembourg/LNS0105586/2020\|EPI_ISL_739713\| | ···················· | ···················· | ····················· | ···················· | ···················· | ···················· |
| Z | 1295 | hCoV-19/Luxembourg/LNS0108329/2020\|EPI_ISL_739714\| | ···················· | ···················· | ····················· | ···················· | ···················· | ···················· |
| Z | 1296 | hCoV-19/Luxembourg/LNS0109742/2020\|EPI_ISL_739715\| | ···················· | ···················· | ····················· | ···················· | ···················· | ···················· |
| Z | 1297 | hCoV-19/Luxembourg/LNS0116884/2020\|EPI_ISL_739716\| | ···················· | ···················· | ····················· | ···················· | ···················· | ···················· |
| Z | 1298 | hCoV-19/Luxembourg/LNS0120826/2020\|EPI_ISL_739717\| | ···················· | ···················· | ····················· | ···················· | ···················· | ···················· |
| Z | 1299 | hCoV-19/Luxembourg/LNS0121983/2020\|EPI_ISL_739718\| | ···················· | ···················· | ····················· | ···················· | ···················· | ···················· |
| Z | 1300 | hCoV-19/Luxembourg/LNS0128091/2020\|EPI_ISL_739719\| | ···················· | ···················· | ····················· | ···················· | ···················· | ···················· |
| Z | 1301 | hCoV-19/Luxembourg/LNS0129194/2020\|EPI_ISL_739720\| | ···················· | ···················· | ····················· | ···················· | ···················· | ···················· |
| Z | 1302 | hCoV-19/Luxembourg/LNS0140663/2020\|EPI_ISL_739721\| | ···················· | ···················· | ····················· | ···················· | ···················· | ···················· |
| Z | 1303 | hCoV-19/Luxembourg/LNS0143745/2020\|EPI_ISL_739722\| | ···················· | ···················· | ····················· | ···················· | ···················· | ···················· |
| Z | 1304 | hCoV-19/Luxembourg/LNS0150051/2020\|EPI_ISL_739723\| | ···················· | ···················· | ····················· | ···················· | ···················· | ···················· |
| Z | 1305 | hCoV-19/Luxembourg/LNS0154197/2020\|EPI_ISL_739724\| | ···················· | ···················· | ····················· | ···················· | ···················· | ···················· |
| Z | 1306 | hCoV-19/Luxembourg/LNS0155076/2020\|EPI_ISL_739725\| | ···················· | ···················· | ····················· | ···················· | ···················· | ···················· |
| Z | 1307 | hCoV-19/Luxembourg/LNS0156004/2020\|EPI_ISL_739726\| | ···················· | ···················· | ····················· | ···················· | ······T············· | ···················· |
| Z | 1308 | hCoV-19/Luxembourg/LNS0172206/2020\|EPI_ISL_739728\| | ···················· | ···················· | ····················· | ···················· | ···················· | ···················· |
| Z | 1309 | hCoV-19/Luxembourg/LNS0175801/2020\|EPI_ISL_739729\| | ···················· | ···················· | ····················· | ···················· | ···················· | ···················· |
| Z | 1310 | hCoV-19/Luxembourg/LNS0181071/2020\|EPI_ISL_739730\| | ···················· | ···················· | ····················· | ···················· | ···················· | ···················· |
| Z | 1311 | hCoV-19/Luxembourg/LNS0181724/2020\|EPI_ISL_739731\| | ···················· | ···················· | ····················· | ···················· | ···················· | ···················· |
| Z | 1312 | hCoV-19/Luxembourg/LNS0184878/2020\|EPI_ISL_739732\| | ···················· | ···················· | ····················· | ···················· | ···················· | ···················· |
| Z | 1313 | hCoV-19/Luxembourg/LNS0186395/2020\|EPI_ISL_739733\| | ···················· | ···················· | ····················· | ···················· | ···················· | ···················· |
| Z | 1314 | hCoV-19/Luxembourg/LNS0187666/2020\|EPI_ISL_739734\| | ···················· | ···················· | ····················· | ···················· | ···················· | ···················· |
| Z | 1315 | hCoV-19/Luxembourg/LNS0194872/2020\|EPI_ISL_739735\| | ···················· | ···················· | ····················· | ···················· | ···················· | ···················· |
| Z | 1316 | hCoV-19/Luxembourg/LNS0205443/2020\|EPI_ISL_739736\| | ···················· | ···················· | ····················· | ···················· | ···················· | ···················· |
| Z | 1317 | hCoV-19/Luxembourg/LNS0209461/2020\|EPI_ISL_739737\| | ···················· | ···················· | ····················· | ···················· | ···················· | ···················· |
| Z | 1318 | hCoV-19/Luxembourg/LNS0215436/2020\|EPI_ISL_739738\| | ···················· | ···················· | ····················· | ···················· | ···················· | ···················· |
| Z | 1319 | hCoV-19/Luxembourg/LNS0224445/2020\|EPI_ISL_739739\| | ···················· | ···················· | ····················· | ···················· | ···················· | ···················· |
| Z | 1320 | hCoV-19/Luxembourg/LNS0224816/2020\|EPI_ISL_739740\| | ···················· | ···················· | ····················· | ···················· | ···················· | ···················· |
| Z | 1321 | hCoV-19/Luxembourg/LNS0225895/2020\|EPI_ISL_739741\| | ···················· | ···················· | ····················· | ···················· | ···················· | ···················· |
| Z | 1322 | hCoV-19/Luxembourg/LNS0247915/2020\|EPI_ISL_739742\| | ···················· | ···················· | ···········T········· | ···················· | ···················· | ···················· |
| Z | 1323 | hCoV-19/Luxembourg/LNS0254753/2020\|EPI_ISL_739743\| | ···················· | ···················· | ····················· | ···················· | ···················· | ···················· |
| Z | 1324 | hCoV-19/Luxembourg/LNS0254853/2020\|EPI_ISL_739744\| | ···················· | ···················· | ····················· | ···················· | ···················· | ···················· |
| Z | 1326 | hCoV-19/Luxembourg/LNS0280372/2020\|EPI_ISL_739748\| | ···················· | ···················· | ····················· | ···················· | ······T············· | ···················· |

| | | | | | | | | |
|---|---|---|---|---|---|---|---|---|
| Z | 1327 | hCoV-19/Luxembourg/LNS0281498/2020\|EPI_ISL_739749\| | ···················· | ···················· | ····················· | ···················· | ···················· | ···················· |
| Z | 1328 | hCoV-19/Luxembourg/LNS0284262/2020\|EPI_ISL_739750\| | ···················· | ···················· | ····················· | ···················· | ···················· | ···················· |
| Z | 1329 | hCoV-19/Luxembourg/LNS0293199/2020\|EPI_ISL_739751\| | ···················· | ···················· | ····················· | ···················· | ···················· | ···················· |
| Z | 1330 | hCoV-19/Luxembourg/LNS0302752/2020\|EPI_ISL_739752\| | ···················· | ···················· | ····················· | ···················· | ···················· | ···················· |
| Z | 1331 | hCoV-19/Luxembourg/LNS0303589/2020\|EPI_ISL_739753\| | ···················· | ···················· | ····················· | ···················· | ···················· | ···················· |
| Z | 1332 | hCoV-19/Luxembourg/LNS0310565/2020\|EPI_ISL_739754\| | ···················· | ···················· | ····················· | ···················· | ···················· | ···················· |
| Z | 1333 | hCoV-19/Luxembourg/LNS0317136/2020\|EPI_ISL_739755\| | ···················· | ···················· | ····················· | ···················· | ···················· | ···················· |
| Z | 1334 | hCoV-19/Luxembourg/LNS0320605/2020\|EPI_ISL_739756\| | ············T······· | ···················· | ····················· | ···················· | ···················· | ···················· |
| Z | 1335 | hCoV-19/Luxembourg/LNS0325922/2020\|EPI_ISL_739757\| | ············T······· | ···················· | ····················· | ···················· | ···················· | ···················· |
| Z | 1336 | hCoV-19/Luxembourg/LNS0333768/2020\|EPI_ISL_739759\| | ···················· | ···················· | ····················· | ···················· | ···················· | ···················· |
| Z | 1337 | hCoV-19/Luxembourg/LNS0438295/2020\|EPI_ISL_739775\| | ···················· | ···················· | ····················· | ···················· | ···················· | ···················· |
| Z | 1338 | hCoV-19/Luxembourg/LNS0454637/2020\|EPI_ISL_739777\| | ···················· | ···················· | ····················· | ···················· | ···················· | ···················· |
| Z | 1339 | hCoV-19/Luxembourg/LNS0459171/2020\|EPI_ISL_739778\| | ···················· | ···················· | ····················· | ···················· | ···················· | ···················· |
| Z | 1340 | hCoV-19/Luxembourg/LNS0462253/2020\|EPI_ISL_739779\| | ···················· | ···················· | ····················· | ···················· | ···················· | ···················· |
| Z | 1341 | hCoV-19/Luxembourg/LNS0468115/2020\|EPI_ISL_739780\| | ···················· | ···················· | ····················· | ···················· | ···················· | ···················· |
| Z | 1342 | hCoV-19/Luxembourg/LNS0469996/2020\|EPI_ISL_739781\| | ···················· | ···················· | ····················· | ···················· | ···················· | ···················· |
| Z | 1343 | hCoV-19/Luxembourg/LNS0470029/2020\|EPI_ISL_739782\| | ···················· | ···················· | ····················· | ···················· | ···················· | ···················· |
| Z | 1344 | hCoV-19/Luxembourg/LNS0478039/2020\|EPI_ISL_739783\| | ···················· | ···················· | ····················· | ···················· | ···················· | ···················· |
| Z | 1345 | hCoV-19/Luxembourg/LNS0479136/2020\|EPI_ISL_739784\| | ···················· | ···················· | ····················· | ···················· | ···················· | ···················· |
| Z | 1346 | hCoV-19/Luxembourg/LNS0483937/2020\|EPI_ISL_739786\| | ···················· | ···················· | ····················· | ···················· | ···················· | ···················· |
| Z | 1347 | hCoV-19/Luxembourg/LNS0490194/2020\|EPI_ISL_739787\| | ···················· | ···················· | ····················· | ···················· | ···················· | ···················· |
| Z | 1348 | hCoV-19/Luxembourg/LNS0492420/2020\|EPI_ISL_739788\| | ···················· | ···················· | ····················· | ···················· | ···················· | ···················· |
| Z | 1349 | hCoV-19/Luxembourg/LNS0493779/2020\|EPI_ISL_739790\| | ···················· | ···················· | ····················· | ···················· | ···················· | ···················· |
| Z | 1350 | hCoV-19/Luxembourg/LNS0497696/2020\|EPI_ISL_739791\| | ···················· | ···················· | ····················· | ···················· | ···················· | ···················· |
| Z | 1351 | hCoV-19/Luxembourg/LNS0499266/2020\|EPI_ISL_739792\| | ···················· | ···················· | ····················· | ···················· | ···················· | ···················· |
| Z | 1352 | hCoV-19/Luxembourg/LNS0507361/2020\|EPI_ISL_739793\| | ···················· | ···················· | ····················· | ···················· | ···················· | ···················· |
| Z | 1353 | hCoV-19/Luxembourg/LNS0511956/2020\|EPI_ISL_739794\| | ···················· | G··················· | ····················· | ···················· | ···················· | ···················· |
| Z | 1354 | hCoV-19/Luxembourg/LNS0519497/2020\|EPI_ISL_739795\| | ···················· | ···················· | ····················· | ···················· | ···················· | ···················· |
| Z | 1355 | hCoV-19/Luxembourg/LNS0553301/2020\|EPI_ISL_739797\| | ···················· | ···················· | ····················· | ···················· | ···················· | ···················· |
| Z | 1356 | hCoV-19/Luxembourg/LNS0554561/2020\|EPI_ISL_739798\| | ···················· | ···················· | ····················· | ···················· | ···················· | ···················· |
| Z | 1357 | hCoV-19/Luxembourg/LNS0566366/2020\|EPI_ISL_739799\| | ···················· | ···················· | ····················· | ···················· | ······T············· | ···················· |
| Z | 1358 | hCoV-19/Canada/BC-BCCDC-7532/2020\|EPI_ISL_740700\|2 | ···················· | ···················· | ····················· | ···················· | ···················· | ···················· |
| Z | 1359 | hCoV-19/Canada/BC-BCCDC-7534/2020\|EPI_ISL_740701\|2 | ···················· | ···················· | ····················· | ···················· | ···················· | ···················· |
| Z | 1360 | hCoV-19/Canada/BC-BCCDC-7535/2020\|EPI_ISL_740702\|2 | ···················· | ···················· | ····················· | ···················· | ···················· | ···················· |
| Z | 1361 | hCoV-19/Canada/BC-BCCDC-7536/2020\|EPI_ISL_740703\|2 | ···················· | ···················· | ····················· | ···················· | ···················· | ···················· |
| Z | 1362 | hCoV-19/Canada/BC-BCCDC-7537/2020\|EPI_ISL_740704\|2 | ···················· | ···················· | ····················· | ···················· | ···················· | ···················· |
| Z | 1363 | hCoV-19/Canada/BC-BCCDC-7538/2020\|EPI_ISL_740705\|2 | ···················· | ···················· | ····················· | ···················· | ···················· | ···················· |
| Z | 1364 | hCoV-19/Canada/BC-BCCDC-7540/2020\|EPI_ISL_740706\|2 | ···················· | ···················· | ····················· | ···················· | ···················· | ···················· |
| Z | 1365 | hCoV-19/Canada/BC-BCCDC-7544/2020\|EPI_ISL_740707\|2 | ···················· | ···················· | ····················· | ···················· | ···················· | ···················· |
| Z | 1366 | hCoV-19/Canada/BC-BCCDC-7545/2020\|EPI_ISL_740708\|2 | ···················· | ···················· | ····················· | ···················· | ···················· | ···················· |
| Z | 1367 | hCoV-19/Canada/BC-BCCDC-7546/2020\|EPI_ISL_740709\|2 | ···················· | ···················· | ····················· | ···················· | ···················· | ···················· |
| Z | 1368 | hCoV-19/Canada/BC-BCCDC-7547/2020\|EPI_ISL_740710\|2 | ···················· | ·A·················· | ····················· | ···················· | ···················· | ···A················ |
| Z | 1369 | hCoV-19/Canada/BC-BCCDC-7548/2020\|EPI_ISL_740711\|2 | ···················· | ···················· | ····················· | ···················· | ···················· | ···················· |
| Z | 1370 | hCoV-19/Canada/BC-BCCDC-7550/2020\|EPI_ISL_740713\|2 | ···················· | ·A·················· | ····················· | ···················· | ···················· | ···A················ |
| Z | 1371 | hCoV-19/Canada/BC-BCCDC-7552/2020\|EPI_ISL_740715\|2 | ···················· | ·A·················· | ····················· | ···················· | ···················· | ···A················ |
| Z | 1372 | hCoV-19/Canada/BC-BCCDC-7555/2020\|EPI_ISL_740717\|2 | ···················· | ·A·················· | ····················· | ···················· | ···················· | ···A················ |
| Z | 1373 | hCoV-19/Canada/BC-BCCDC-7556/2020\|EPI_ISL_740718\|2 | ···················· | ···················· | ····················· | ···················· | ···················· | ···················· |
| Z | 1374 | hCoV-19/Canada/BC-BCCDC-7557/2020\|EPI_ISL_740719\|2 | ···················· | ·A·················· | ····················· | ···················· | ···················· | ···A················ |
| Z | 1375 | hCoV-19/Canada/BC-BCCDC-7558/2020\|EPI_ISL_740720\|2 | ···················· | ···················· | ····················· | ···················· | ···················· | ···················· |
| Z | 1376 | hCoV-19/Canada/BC-BCCDC-7559/2020\|EPI_ISL_740721\|2 | ···················· | ···················· | ····················· | ···················· | ···················· | ···················· |
| Z | 1377 | hCoV-19/Canada/BC-BCCDC-7562/2020\|EPI_ISL_740723\|2 | ···················· | ···················· | ····················· | ···················· | ···················· | ···················· |
| Z | 1378 | hCoV-19/Canada/BC-BCCDC-7563/2020\|EPI_ISL_740724\|2 | ···················· | ·A·················· | ····················· | ···················· | ···················· | ···A················ |
| Z | 1379 | hCoV-19/Canada/BC-BCCDC-7566/2020\|EPI_ISL_740727\|2 | ···················· | ···················· | ····················· | ···················· | ···················· | ···················· |
| Z | 1380 | hCoV-19/Canada/BC-BCCDC-7569/2020\|EPI_ISL_740728\|2 | ···················· | ···················· | ····················· | ···················· | ···················· | ···················· |
| Z | 1381 | hCoV-19/Canada/BC-BCCDC-7574/2020\|EPI_ISL_740729\|2 | ···················· | ···················· | ····················· | ···················· | ···················· | ···················· |
| Z | 1382 | hCoV-19/Canada/BC-BCCDC-7575/2020\|EPI_ISL_740730\|2 | ···················· | ···················· | ····················· | ···················· | ···················· | ···················· |
| Z | 1383 | hCoV-19/Canada/BC-BCCDC-7576/2020\|EPI_ISL_740731\|2 | ···················· | ·A·················· | ····················· | ···················· | ···················· | ···A················ |
| Z | 1384 | hCoV-19/Canada/BC-BCCDC-7587/2020\|EPI_ISL_740735\|2 | ···················· | ·A·················· | ····················· | ···················· | ···················· | ···A················ |
| Z | 1385 | hCoV-19/Canada/BC-BCCDC-7588/2020\|EPI_ISL_740736\|2 | ···················· | ·A·················· | ····················· | ···················· | ···················· | ···A················ |
| Z | 1386 | hCoV-19/Canada/BC-BCCDC-7591/2020\|EPI_ISL_740738\|2 | ···················· | ···················· | ····················· | ···················· | ···················· | ···················· |
| Z | 1387 | hCoV-19/Canada/BC-BCCDC-7592/2020\|EPI_ISL_740739\|2 | ···················· | ···················· | ····················· | ···················· | ···················· | ···················· |
| Z | 1388 | hCoV-19/Canada/BC-BCCDC-7593/2020\|EPI_ISL_740740\|2 | ···················· | ·A·················· | ····················· | ···················· | ···················· | ···A················ |
| Z | 1389 | hCoV-19/Canada/BC-BCCDC-8363/2020\|EPI_ISL_740753\|2 | ···················· | ···················· | ····················· | ···················· | ···················· | ···················· |
| Z | 1390 | hCoV-19/Canada/BC-BCCDC-8364/2020\|EPI_ISL_740754\|2 | ···················· | ···················· | ····················· | ···················· | ···················· | ···················· |
| Z | 1391 | hCoV-19/Canada/BC-BCCDC-8371/2020\|EPI_ISL_740760\|2 | ···················· | ·A·················· | ····················· | ···················· | ···················· | ···A················ |
| Z | 1392 | hCoV-19/Canada/BC-BCCDC-8375/2020\|EPI_ISL_740761\|2 | ···················· | ···················· | ····················· | ···················· | ···················· | ···················· |
| Z | 1393 | hCoV-19/Canada/BC-BCCDC-8377/2020\|EPI_ISL_740762\|2 | ···················· | ···················· | ····················· | ···················· | ···················· | ···················· |
| Z | 1394 | hCoV-19/Canada/BC-BCCDC-8379/2020\|EPI_ISL_740763\|2 | ···················· | ···················· | ····················· | ···················· | ···················· | ···················· |
| Z | 1395 | hCoV-19/Canada/BC-BCCDC-8380/2020\|EPI_ISL_740764\|2 | ···················· | ·A·················· | ····················· | ···················· | ···················· | ···A················ |
| Z | 1396 | hCoV-19/Canada/BC-BCCDC-8381/2020\|EPI_ISL_740765\|2 | ···················· | ···················· | ····················· | ···················· | ···················· | ···················· |
| Z | 1397 | hCoV-19/Canada/BC-BCCDC-8384/2020\|EPI_ISL_740766\|2 | ···················· | ·A·················· | ····················· | ···················· | ···················· | ···A················ |
| Z | 1398 | hCoV-19/Canada/BC-BCCDC-8387/2020\|EPI_ISL_740767\|2 | ···················· | ···················· | ····················· | ···················· | ···················· | ···················· |
| Z | 1399 | hCoV-19/Canada/BC-BCCDC-8388/2020\|EPI_ISL_740768\|2 | ···················· | ·A·················· | ····················· | ···················· | ···················· | ···A················ |
| Z | 1400 | hCoV-19/Canada/BC-BCCDC-8392/2020\|EPI_ISL_740769\|2 | ···················· | ·A·················· | ····················· | ···················· | ···················· | ···A················ |
| Z | 1401 | hCoV-19/Canada/BC-BCCDC-8393/2020\|EPI_ISL_740770\|2 | ···················· | ···················· | ····················· | ···················· | ···················· | ···················· |
| Z | 1402 | hCoV-19/Canada/BC-BCCDC-8394/2020\|EPI_ISL_740771\|2 | ···················· | ·A·················· | ····················· | ···················· | ···················· | ···A················ |
| Z | 1403 | hCoV-19/Canada/BC-BCCDC-8397/2020\|EPI_ISL_740773\|2 | ···················· | ···················· | ····················· | ···················· | ···················· | ···················· |
| Z | 1404 | hCoV-19/Canada/BC-BCCDC-8398/2020\|EPI_ISL_740774\|2 | ···················· | ···················· | ····················· | ···················· | ···················· | ···················· |
| Z | 1405 | hCoV-19/Canada/BC-BCCDC-8399/2020\|EPI_ISL_740775\|2 | ···················· | ···················· | ····················· | ···················· | ···················· | ···················· |
| Z | 1406 | hCoV-19/Canada/BC-BCCDC-8406/2020\|EPI_ISL_740778\|2 | ···················· | ···················· | ····················· | ···················· | ···················· | ···················· |

| | | | | | | | | |
|---|---|---|---|---|---|---|---|---|
| Z | 1407 | hCoV-19/Canada/BC-BCCDC-8407/2020\|EPI_ISL_740779\|2 | ···················· | ···················· | ···················· | ···················· | ···················· | ···················· |
| Z | 1408 | hCoV-19/Canada/BC-BCCDC-8409/2020\|EPI_ISL_740781\|2 | ···················· | ·A·················· | ···················· | ···················· | ···················· | ···A················ |
| Z | 1409 | hCoV-19/Canada/BC-BCCDC-8410/2020\|EPI_ISL_740782\|2 | ···················· | ···················· | ···················· | ···················· | ···················· | ···················· |
| Z | 1410 | hCoV-19/Canada/BC-BCCDC-8411/2020\|EPI_ISL_740783\|2 | ···················· | ·A·················· | ···················· | ···················· | ···················· | ···A················ |
| Z | 1411 | hCoV-19/Canada/BC-BCCDC-8412/2020\|EPI_ISL_740784\|2 | ···················· | ···················· | ···················· | ···················· | ···················· | ···················· |
| Z | 1412 | hCoV-19/Canada/BC-BCCDC-8413/2020\|EPI_ISL_740785\|2 | ···················· | ·A·················· | ···················· | ···················· | ···················· | ···A················ |
| Z | 1413 | hCoV-19/Luxembourg/LNS5203949/2020\|EPI_ISL_744201\| | ···················· | ···················· | ···················· | ···················· | ···················· | ···················· |
| Z | 1414 | hCoV-19/Luxembourg/LNS5215473/2020\|EPI_ISL_744202\| | ···················· | ···················· | ···················· | ···················· | ···················· | ···················· |
| Z | 1415 | hCoV-19/Luxembourg/LNS5217472/2020\|EPI_ISL_744203\| | ···················· | ···················· | ···················· | ···················· | ···················· | ···················· |
| Z | 1416 | hCoV-19/Luxembourg/LNS5227811/2020\|EPI_ISL_744204\| | ···················· | ···················· | ···················· | ···················· | ···················· | ···················· |
| Z | 1417 | hCoV-19/Luxembourg/LNS5232659/2020\|EPI_ISL_744205\| | ···················· | ···················· | ···················· | ···················· | ···················· | ···················· |
| Z | 1418 | hCoV-19/Luxembourg/LNS5234553/2020\|EPI_ISL_744206\| | ···················· | ···················· | ···················· | ···················· | ···················· | ···················· |
| Z | 1419 | hCoV-19/Luxembourg/LNS5238472/2020\|EPI_ISL_744207\| | ···················· | ···················· | ···················· | ···················· | ···················· | ···················· |
| Z | 1420 | hCoV-19/Luxembourg/LNS5242152/2020\|EPI_ISL_744208\| | ···················· | ···················· | ···················· | ···················· | ···················· | ···················· |
| Z | 1421 | hCoV-19/Luxembourg/LNS5256710/2020\|EPI_ISL_744209\| | ···················· | G··················· | ···················· | ···················· | ···················· | ···················· |
| Z | 1422 | hCoV-19/Luxembourg/LNS5262261/2020\|EPI_ISL_744210\| | ···················· | ···················· | ···················· | ···················· | ···················· | ···················· |
| Z | 1423 | hCoV-19/Luxembourg/LNS5279966/2020\|EPI_ISL_744212\| | ···················· | ···················· | ···················· | ···················· | ···················· | ···················· |
| Z | 1424 | hCoV-19/Luxembourg/LNS5301713/2020\|EPI_ISL_744213\| | ···················· | ···················· | ···················· | ···················· | ···················· | ···················· |
| Z | 1425 | hCoV-19/Luxembourg/LNS5304019/2020\|EPI_ISL_744214\| | ···················· | ···················· | ···················· | ···················· | ···················· | ···················· |
| Z | 1426 | hCoV-19/Luxembourg/LNS5315916/2020\|EPI_ISL_744215\| | ···················· | ···················· | ···················· | ···················· | ···················· | ···················· |
| Z | 1427 | hCoV-19/Luxembourg/LNS5317025/2020\|EPI_ISL_744216\| | ···················· | ···················· | ···················· | ···················· | ···················· | ···················· |
| Z | 1428 | hCoV-19/Luxembourg/LNS5322549/2020\|EPI_ISL_744217\| | ···················· | ···················· | ···················· | ···················· | ······T············· | ···················· |
| Z | 1429 | hCoV-19/Luxembourg/LNS5328839/2020\|EPI_ISL_744218\| | ···················· | ···················· | ···················· | ···················· | ···················· | ···················· |
| Z | 1430 | hCoV-19/Luxembourg/LNS5329793/2020\|EPI_ISL_744219\| | ···················· | ···················· | ···················· | ···················· | ···················· | ···················· |
| Z | 1431 | hCoV-19/Luxembourg/LNS5339140/2020\|EPI_ISL_744220\| | ···················· | ···················· | ···················· | ···················· | ···················· | ···················· |
| Z | 1432 | hCoV-19/Luxembourg/LNS5350981/2020\|EPI_ISL_744221\| | ···················· | ···················· | ···················· | ···················· | ···················· | ···················· |
| Z | 1433 | hCoV-19/Luxembourg/LNS5356686/2020\|EPI_ISL_744222\| | ···················· | ···················· | ···················· | ···················· | ···················· | ···················· |
| Z | 1434 | hCoV-19/Luxembourg/LNS5364228/2020\|EPI_ISL_744223\| | ···················· | ···················· | ···················· | ···················· | ···················· | ···················· |
| Z | 1435 | hCoV-19/Luxembourg/LNS5381060/2020\|EPI_ISL_744224\| | ···················· | ···················· | ···················· | ···················· | ···················· | ···················· |
| Z | 1436 | hCoV-19/Luxembourg/LNS5395834/2020\|EPI_ISL_744225\| | ···················· | ···················· | ···················· | ···················· | ···················· | ···················· |
| Z | 1437 | hCoV-19/Luxembourg/LNS5396868/2020\|EPI_ISL_744226\| | ···················· | ···················· | ···················· | ···················· | ···················· | ···················· |
| Z | 1438 | hCoV-19/Luxembourg/LNS5406700/2020\|EPI_ISL_744227\| | ···················· | ···················· | ···················· | ···················· | ···················· | ···················· |
| Z | 1439 | hCoV-19/Luxembourg/LNS5415838/2020\|EPI_ISL_744228\| | ···················· | ···················· | ···················· | ···················· | ···················· | ···················· |
| Z | 1440 | hCoV-19/Luxembourg/LNS5416298/2020\|EPI_ISL_744229\| | ···················· | ···················· | ···················· | ···················· | ···················· | ···················· |
| Z | 1441 | hCoV-19/Luxembourg/LNS5416537/2020\|EPI_ISL_744230\| | ···················· | ···················· | ···················· | ···················· | ···················· | ···················· |
| Z | 1442 | hCoV-19/Luxembourg/LNS5450477/2020\|EPI_ISL_744231\| | ···················· | ···················· | ···················· | ···················· | ···················· | ···················· |
| Z | 1443 | hCoV-19/Luxembourg/LNS5454479/2020\|EPI_ISL_744232\| | ···················· | ···················· | ···················· | ···················· | ···················· | ···················· |
| Z | 1444 | hCoV-19/Luxembourg/LNS5454756/2020\|EPI_ISL_744233\| | ···················· | ···················· | ···················· | ···················· | ···················· | ···················· |
| Z | 1445 | hCoV-19/Luxembourg/LNS5457256/2020\|EPI_ISL_744234\| | ···················· | ···················· | ···················· | ···················· | ···················· | ···················· |
| Z | 1446 | hCoV-19/Luxembourg/LNS5457824/2020\|EPI_ISL_744235\| | ···················· | ···················· | ···················· | ···················· | ···················· | ···················· |
| Z | 1447 | hCoV-19/Luxembourg/LNS5462232/2020\|EPI_ISL_744236\| | ···················· | G··················· | ···················· | ···················· | ···················· | ···················· |
| Z | 1448 | hCoV-19/Luxembourg/LNS5467958/2020\|EPI_ISL_744237\| | ···················· | ···················· | ···················· | ···················· | ···················· | ···················· |
| Z | 1449 | hCoV-19/Luxembourg/LNS5473237/2020\|EPI_ISL_744238\| | ···················· | ···················· | ···················· | ···················· | ···················· | ···················· |
| Z | 1450 | hCoV-19/Luxembourg/LNS5474384/2020\|EPI_ISL_744239\| | ···················· | ···················· | ···················· | ···················· | ···················· | ···················· |
| Z | 1451 | hCoV-19/Luxembourg/LNS5490337/2020\|EPI_ISL_744240\| | ···················· | ···················· | ···················· | ···················· | ···················· | ···················· |
| Z | 1452 | hCoV-19/Luxembourg/LNS5491473/2020\|EPI_ISL_744241\| | ···················· | ···················· | ···················· | ···················· | ······T············· | ···················· |
| Z | 1453 | hCoV-19/Luxembourg/LNS5493972/2020\|EPI_ISL_744242\| | ···················· | ····A··············· | ···················· | ···················· | ···················· | ···················· |
| Z | 1454 | hCoV-19/Luxembourg/LNS5506489/2020\|EPI_ISL_744243\| | ···················· | ···················· | ···················· | ···················· | ···················· | ···················· |
| Z | 1455 | hCoV-19/Luxembourg/LNS5509035/2020\|EPI_ISL_744244\| | ···················· | ···················· | ···················· | ···················· | ···················· | ···················· |
| Z | 1456 | hCoV-19/Luxembourg/LNS5518532/2020\|EPI_ISL_744245\| | ···················· | ···················· | ···················· | ···················· | ···················· | ···················· |
| Z | 1457 | hCoV-19/Luxembourg/LNS5523733/2020\|EPI_ISL_744246\| | ···················· | ···················· | ···················· | ···················· | ···C················ | ···················· |
| Z | 1458 | hCoV-19/Luxembourg/LNS5532570/2020\|EPI_ISL_744247\| | ···················· | ···················· | ···················· | ···················· | ···················· | ···················· |
| Z | 1459 | hCoV-19/Luxembourg/LNS5539267/2020\|EPI_ISL_744248\| | ···················· | ···················· | ···················· | ···················· | ···················· | ···················· |
| Z | 1460 | hCoV-19/Luxembourg/LNS5558475/2020\|EPI_ISL_744250\| | ···················· | ···················· | ···················· | ···················· | ···················· | ···················· |
| Z | 1461 | hCoV-19/Luxembourg/LNS5559609/2020\|EPI_ISL_744251\| | ···················· | ···················· | ···················· | ···················· | ···················· | ···················· |
| Z | 1462 | hCoV-19/Luxembourg/LNS5559819/2020\|EPI_ISL_744252\| | ···················· | ···················· | ···················· | ···················· | ···················· | ···················· |
| Z | 1463 | hCoV-19/Luxembourg/LNS5561396/2020\|EPI_ISL_744253\| | ···················· | ···················· | ···················· | ···················· | ···················· | ···················· |
| Z | 1464 | hCoV-19/Luxembourg/LNS5578604/2020\|EPI_ISL_744254\| | ···················· | ···················· | ···················· | ···················· | ···················· | ···················· |
| Z | 1465 | hCoV-19/Luxembourg/LNS5579246/2020\|EPI_ISL_744255\| | ···················· | G··················· | ···················· | ···················· | ···················· | ···················· |
| Z | 1466 | hCoV-19/Luxembourg/LNS5580084/2020\|EPI_ISL_744256\| | ···················· | ···················· | ···················· | ···················· | ···················· | ···················· |
| Z | 1467 | hCoV-19/Luxembourg/LNS5591357/2020\|EPI_ISL_744257\| | ···················· | ···················· | ···················· | ···················· | ···················· | ···················· |
| Z | 1468 | hCoV-19/Luxembourg/LNS5595068/2020\|EPI_ISL_744258\| | ···················· | ···················· | ···················· | ···················· | ···················· | ···················· |
| Z | 1469 | hCoV-19/Luxembourg/LNS5605056/2020\|EPI_ISL_744260\| | ···················· | ···················· | ···················· | ···················· | ···················· | ···················· |
| Z | 1470 | hCoV-19/Luxembourg/LNS5621871/2020\|EPI_ISL_744261\| | ···T················ | ···················· | ···················· | ···················· | ···················· | ···················· |
| Z | 1471 | hCoV-19/Luxembourg/LNS5622303/2020\|EPI_ISL_744262\| | ···················· | ···················· | ···················· | ···················· | ···················· | ···················· |
| Z | 1472 | hCoV-19/Luxembourg/LNS5628764/2020\|EPI_ISL_744263\| | ···················· | ···················· | ···················· | ···················· | ······T············· | ···················· |
| Z | 1473 | hCoV-19/Luxembourg/LNS5632061/2020\|EPI_ISL_744264\| | ···················· | ···················· | ···················· | ···················· | ···················· | ···················· |
| Z | 1474 | hCoV-19/Luxembourg/LNS5632110/2020\|EPI_ISL_744265\| | ···················· | ···················· | ···················· | ···················· | ···················· | ···················· |
| Z | 1475 | hCoV-19/Luxembourg/LNS5636946/2020\|EPI_ISL_744266\| | ···················· | ···················· | ···················· | ···················· | ···················· | ···················· |
| Z | 1476 | hCoV-19/Luxembourg/LNS5638426/2020\|EPI_ISL_744267\| | ···················· | ···················· | ···················· | ···················· | ···················· | ···················· |
| Z | 1477 | hCoV-19/Luxembourg/LNS5650133/2020\|EPI_ISL_744268\| | ···················· | ···················· | ···················· | ···················· | ···················· | ···················· |
| Z | 1478 | hCoV-19/Luxembourg/LNS5663351/2020\|EPI_ISL_744270\| | ···················· | ···················· | ···················· | ···················· | ······T············· | ···················· |
| Z | 1479 | hCoV-19/Luxembourg/LNS5675141/2020\|EPI_ISL_744271\| | ···················· | G··················· | ···················· | ···················· | ···················· | ···················· |
| Z | 1480 | hCoV-19/Luxembourg/LNS5677307/2020\|EPI_ISL_744272\| | ···················· | ···················· | ···················· | ···················· | ···················· | ···················· |
| Z | 1481 | hCoV-19/Luxembourg/LNS5684173/2020\|EPI_ISL_744273\| | ···················· | ···················· | ···················· | ···················· | ···················· | ···················· |
| Z | 1482 | hCoV-19/Luxembourg/LNS5694108/2020\|EPI_ISL_744275\| | ···················· | ···················· | ···················· | ···················· | ···················· | ···················· |
| Z | 1483 | hCoV-19/Luxembourg/LNS5694717/2020\|EPI_ISL_744276\| | ···················· | ···················· | ···················· | ···················· | ···················· | ···················· |
| Z | 1484 | hCoV-19/Luxembourg/LNS5699520/2020\|EPI_ISL_744277\| | ···················· | G··················· | ···················· | ···················· | ···················· | ···················· |
| Z | 1485 | hCoV-19/Luxembourg/LNS5702473/2020\|EPI_ISL_744278\| | ···················· | ···················· | ···················· | ···················· | ···················· | ···················· |
| Z | 1486 | hCoV-19/Luxembourg/LNS5704143/2020\|EPI_ISL_744280\| | ···················· | ···················· | ···················· | ···················· | ···················· | ···················· |

Z 1487 hCoV-19/Luxembourg/LNS5706546/2020|EPI_ISL_744281| ···················· ···················· ···················· ···················· ···················· ····················
Z 1488 hCoV-19/Luxembourg/LNS5706571/2020|EPI_ISL_744282| ···················· ···················· ···················· ···················· ···················· ····················
Z 1489 hCoV-19/Luxembourg/LNS5711580/2020|EPI_ISL_744283| ···················· ···················· ···················· ···················· ···················· ····················
Z 1490 hCoV-19/Luxembourg/LNS5718931/2020|EPI_ISL_744284| ···················· ···················· ···················· ···················· ···················· ····················
Z 1491 hCoV-19/Luxembourg/LNS5720760/2020|EPI_ISL_744285| ···················· ···················· ···················· ···················· ···················· ····················
Z 1492 hCoV-19/Luxembourg/LNS5726689/2020|EPI_ISL_744286| ···················· ···················· ···················· ···················· ···················· ····················
Z 1493 hCoV-19/Luxembourg/LNS5734017/2020|EPI_ISL_744287| ···················· ···················· ···················· ···················· ···················· ····················
Z 1494 hCoV-19/Luxembourg/LNS5739154/2020|EPI_ISL_744288| ···················· ···················· ···················· ···················· ···················· ····················
Z 1495 hCoV-19/Luxembourg/LNS5743816/2020|EPI_ISL_744289| ···················· ···················· ···················· ···················· ·····T·············· ····················
Z 1496 hCoV-19/Luxembourg/LNS5744578/2020|EPI_ISL_744290| ···················· ···················· ···················· ···················· ···················· ····················
Z 1497 hCoV-19/Luxembourg/LNS5751723/2020|EPI_ISL_744292| ···················· ···················· ···················· ···················· ···················· ····················
Z 1498 hCoV-19/Luxembourg/LNS5751796/2020|EPI_ISL_744293| ···················· ···················· ···················· ···················· ···················· ····················
Z 1499 hCoV-19/Luxembourg/LNS5753562/2020|EPI_ISL_744294| ···················· ···················· ···················· ···················· ···················· ····················
Z 1500 hCoV-19/Luxembourg/LNS5754655/2020|EPI_ISL_744295| ···················· ···················· ···················· ···················· ···················· ····················
Z 1501 hCoV-19/Luxembourg/LNS5766113/2020|EPI_ISL_744296| ···················· ···················· ···················· ···················· ···················· ····················
Z 1502 hCoV-19/Luxembourg/LNS5766578/2020|EPI_ISL_744297| ···················· ···················· ···················· ···················· ···················· ····················
Z 1503 hCoV-19/Luxembourg/LNS5766990/2020|EPI_ISL_744298| ···················· ···················· ···················· ···················· ···················· ····················
Z 1504 hCoV-19/Luxembourg/LNS5769764/2020|EPI_ISL_744299| ···················· ···················· ···················· ···················· ···················· ····················
Z 1505 hCoV-19/USA/UT-UPHL-2012624197/2020|EPI_ISL_745592 ···················· ···················· ···················· ···················· ···················· ····················
Z 1506 hCoV-19/USA/UT-UPHL-2012294781/2020|EPI_ISL_745593 ···················· ···················· ···················· ···················· ···················· ····················
Z 1507 hCoV-19/USA/UT-UPHL-2012627336/2020|EPI_ISL_745594 ···················· ···················· ···················· ···················· ···················· ····················
Z 1508 hCoV-19/USA/UT-UPHL-2012436582/2020|EPI_ISL_745595 ···················· ···················· ···················· ···················· ···················· ····················
Z 1509 hCoV-19/USA/UT-UPHL-2012308572/2020|EPI_ISL_745596 ···················· ···················· ···················· ···················· ···················· ····················
Z 1510 hCoV-19/USA/UT-UPHL-2012311714/2020|EPI_ISL_745597 ···················· ···················· ···················· ···················· ···················· ····················
Z 1511 hCoV-19/USA/UT-UPHL-2012565545/2020|EPI_ISL_745598 ···················· ···················· ···················· ···················· ···················· ····················
Z 1512 hCoV-19/Chile/RM-84398/2020|EPI_ISL_746803|2020-05 ···················· ···················· ···················· ···················· ···················· ····················
Z 1513 hCoV-19/Chile/RM-84400/2020|EPI_ISL_746804|2020-05 ···················· ···················· ···················· ···················· ···················· ····················
Z 1514 hCoV-19/Chile/RM-84488/2020|EPI_ISL_746805|2020-05 ···················· ···················· ···················· ···················· ···················· ····················
Z 1515 hCoV-19/Chile/RM-84496/2020|EPI_ISL_746806|2020-05 ···················· ···················· ···················· ···················· ···················· ····················
Z 1516 hCoV-19/Chile/RM-84569/2020|EPI_ISL_746807|2020-05 ···················· ···················· ···················· ···················· ···················· ····················
Z 1517 hCoV-19/Chile/RM-84641/2020|EPI_ISL_746808|2020-05 ···················· ···················· ···················· ···················· ···················· ····················
Z 1518 hCoV-19/Chile/RM-84921/2020|EPI_ISL_746809|2020-05 ···················· ···················· ···················· ···················· ···················· ····················
Z 1519 hCoV-19/Chile/RM-84953/2020|EPI_ISL_746810|2020-05 ···················· ···················· ···················· ···················· ···················· ····················
Z 1520 hCoV-19/Chile/RM-85089/2020|EPI_ISL_746811|2020-06 ···················· ···················· ···················· ···················· ···················· ····················
Z 1521 hCoV-19/Chile/RM-85307/2020|EPI_ISL_746812|2020-06 ···················· ···················· ···················· ···················· ···················· ····················
Z 1522 hCoV-19/Chile/RM-85457/2020|EPI_ISL_746813|2020-05 ···················· ···················· ···················· ···················· ···················· ····················
Z 1523 hCoV-19/Chile/RM-85695/2020|EPI_ISL_746814|2020-05 ···················· ···················· ···················· ···················· ···················· ····················
Z 1524 hCoV-19/Chile/RM-86748-TC1/2020|EPI_ISL_746815|202 ···················· ···················· ···················· ···················· ···················· ····················
Z 1525 hCoV-19/Chile/RM-87391-TC1/2020|EPI_ISL_746816|202 ···················· ···················· ···················· ···················· ···················· ····················
Z 1526 hCoV-19/Chile/RM-87410-TC1/2020|EPI_ISL_746817|202 ···················· ···················· ···················· ···················· ···················· ····················
Z 1527 hCoV-19/Chile/RM-87464-TC1/2020|EPI_ISL_746818|202 ···················· ···················· ···················· ···················· ···················· ····················
Z 1528 hCoV-19/Chile/RM-87471-TC1/2020|EPI_ISL_746819|202 ···················· ···················· ···················· ···················· ···················· ····················
Z 1529 hCoV-19/Chile/RM-87477-TC1/2020|EPI_ISL_746820|202 ···················· ···················· ···················· ···················· ···················· ····················
Z 1530 hCoV-19/Chile/RM-87636-TC1/2020|EPI_ISL_746821|202 ···················· ···················· ···················· ···················· ···················· ····················
Z 1531 hCoV-19/Chile/RM-87637-TC1/2020|EPI_ISL_746822|202 ···················· ···················· ···················· ···················· ···················· ····················
Z 1532 hCoV-19/Chile/RM-88024-TC1/2020|EPI_ISL_746823|202 ···················· ···················· ···················· ···················· ···················· ····················
Z 1533 hCoV-19/Chile/RM-88139-TC1/2020|EPI_ISL_746824|202 ···················· ···················· ···················· ···················· ···················· ····················
Z 1534 hCoV-19/USA/UT-UPHL-2012221080/2020|EPI_ISL_746888 ···················· ···················· ···················· ···················· ···················· ····················
Z 1535 hCoV-19/USA/UT-UPHL-2012419639/2020|EPI_ISL_746889 ···················· ···················· ···················· ···················· ···················· ····················
Z 1536 hCoV-19/USA/UT-UPHL-2012147781/2020|EPI_ISL_746890 ···················· ···················· ···················· ···················· ···················· ····················
Z 1537 hCoV-19/Luxembourg/LNS0351170/2020|EPI_ISL_739760| ···················· ···················· ···················· ···················· ···················· ····················
Z 1538 hCoV-19/Luxembourg/LNS0355132/2020|EPI_ISL_739761| ···················· ···················· ···················· ···················· ···················· ····················
Z 1539 hCoV-19/Luxembourg/LNS0355250/2020|EPI_ISL_739762| ···················· ···················· ···················· ···················· ···················· ····················
Z 1540 hCoV-19/Luxembourg/LNS0372015/2020|EPI_ISL_739763| ···················· G··················· ···················· ···················· ···················· ····················
Z 1541 hCoV-19/Luxembourg/LNS0375541/2020|EPI_ISL_739764| ···················· ···················· ···················· ···················· ···················· ····················
Z 1542 hCoV-19/Luxembourg/LNS0375654/2020|EPI_ISL_739765| ···················· ···················· ···················· ···················· ···················· ····················
Z 1543 hCoV-19/Luxembourg/LNS0386826/2020|EPI_ISL_739766| ···················· ···················· ···················· ···················· ···················· ····················
Z 1544 hCoV-19/Luxembourg/LNS0392345/2020|EPI_ISL_739767| ···················· ···················· ···················· ···················· ···················· ····················
Z 1545 hCoV-19/Luxembourg/LNS0394050/2020|EPI_ISL_739768| ···················· G··················· ···················· ···················· ···················· ····················
Z 1546 hCoV-19/Luxembourg/LNS0406231/2020|EPI_ISL_739769| ···················· ···················· ···················· ···················· ···················· ····················
Z 1547 hCoV-19/Luxembourg/LNS0417228/2020|EPI_ISL_739770| ···················· ···················· ···················· ···················· ···················· ····················
Z 1548 hCoV-19/Luxembourg/LNS0418639/2020|EPI_ISL_739771| ···················· ···················· ···················· ···················· ···················· ····················
Z 1549 hCoV-19/Luxembourg/LNS0419896/2020|EPI_ISL_739772| ···················· ···················· ···················· ···················· ···················· ····················
Z 1550 hCoV-19/Luxembourg/LNS0423098/2020|EPI_ISL_739773| ···················· ···················· ···················· ···················· ···················· ····················
Z 1551 hCoV-19/Luxembourg/LNS0434313/2020|EPI_ISL_739774| ···················· ···················· ···················· ···················· ···················· ····················
Z 1552 hCoV-19/Luxembourg/LNS0452279/2020|EPI_ISL_739776| ···················· ···················· ···················· ···················· ···················· ····················
Z 1553 hCoV-19/Canada/BC-BCCDC-7595/2020|EPI_ISL_740742|2 ···················· ···················· ···················· ···················· ···················· ····················
Z 1554 hCoV-19/Canada/BC-BCCDC-7596/2020|EPI_ISL_740743|2 ···················· ···················· ···················· ···················· ···················· ····················
Z 1555 hCoV-19/Canada/BC-BCCDC-7599/2020|EPI_ISL_740744|2 ···················· ···················· ···················· ···················· ···················· ····················
Z 1556 hCoV-19/Canada/BC-BCCDC-7600/2020|EPI_ISL_740745|2 ···················· ···················· ···················· ···················· ···················· ····················
Z 1557 hCoV-19/Canada/BC-BCCDC-7601/2020|EPI_ISL_740746|2 ···················· ·A·················· ···················· ···················· ···················· ··A·················
Z 1558 hCoV-19/Canada/BC-BCCDC-7602/2020|EPI_ISL_740747|2 ···················· ···················· ···················· ···················· ···················· ····················
Z 1559 hCoV-19/Canada/BC-BCCDC-7603/2020|EPI_ISL_740748|2 ···················· ·····A·············· ···················· ···················· ···················· ····················
Z 1560 hCoV-19/Canada/BC-BCCDC-8356/2020|EPI_ISL_740749|2 ···················· ···················· ···················· ···················· ···················· ····················
Z 1561 hCoV-19/Canada/BC-BCCDC-8358/2020|EPI_ISL_740750|2 ···················· ···················· ···················· ···················· ···················· ····················
Z 1562 hCoV-19/Canada/BC-BCCDC-8360/2020|EPI_ISL_740751|2 ···················· ·A·················· ···················· ···················· ···················· ··A·················
Z 1563 hCoV-19/Canada/BC-BCCDC-8362/2020|EPI_ISL_740752|2 ···················· ···················· ···················· ···················· ···················· ····················
Z 1564 hCoV-19/Canada/BC-BCCDC-8366/2020|EPI_ISL_740755|2 ···················· ·A·················· ···················· ···················· ···················· ··A·················
Z 1565 hCoV-19/Canada/BC-BCCDC-8367/2020|EPI_ISL_740756|2 ···················· ···················· ···················· ···················· ···················· ····················
Z 1566 hCoV-19/Canada/BC-BCCDC-8368/2020|EPI_ISL_740757|2 ···················· ···················· ···················· ···················· ···················· ····················

```
Z 1567 hCoV-19/Canada/BC-BCCDC-8369/2020|EPI_ISL_740758|2 ···················· ···················· ···················· ···················· ···················· ····················
Z 1568 hCoV-19/Canada/BC-BCCDC-8370/2020|EPI_ISL_740759|2 ···················· ·A·················· ···················· ···················· ···················· ···A················
Z 1569 hCoV-19/Canada/BC-BCCDC-8418/2020|EPI_ISL_740788|2 ···················· ·A·················· ···················· ···················· ···················· ···A················
Z 1570 hCoV-19/Canada/BC-BCCDC-8420/2020|EPI_ISL_740790|2 ···················· ···················· ···················· ···················· ···················· ····················
Z 1571 hCoV-19/Canada/BC-BCCDC-8422/2020|EPI_ISL_740791|2 ···················· ···················· ···················· ···················· ···················· ····················
Z 1572 hCoV-19/Canada/BC-BCCDC-8425/2020|EPI_ISL_740792|2 ···················· ···················· ···················· ···················· ···················· ····················
Z 1573 hCoV-19/Canada/BC-BCCDC-8426/2020|EPI_ISL_740793|2 ···················· ···················· ···················· ···················· ···················· ····················
Z 1574 hCoV-19/Canada/BC-BCCDC-8429/2020|EPI_ISL_740795|2 ···················· ·A·················· ···················· ···················· ···················· ···A················
Z 1575 hCoV-19/Canada/BC-BCCDC-8430/2020|EPI_ISL_740796|2 ···················· ···················· ···················· ···················· ···················· ····················
Z 1576 hCoV-19/Canada/BC-BCCDC-8431/2020|EPI_ISL_740797|2 ···················· ···················· ···················· ···················· ···················· ····················
Z 1577 hCoV-19/Canada/BC-BCCDC-8433/2020|EPI_ISL_740799|2 ···················· ···················· ···················· ···················· ···················· ····················
Z 1578 hCoV-19/Wales/PHWC-49E407/2020|EPI_ISL_743000|2020 ···················· ···················· ···················· ···················· ···················· ····················
Z 1579 hCoV-19/Wales/PHWC-49E434/2020|EPI_ISL_743001|2020 ···················· ···················· ···················· ···················· ···················· ····················
Z 1580 hCoV-19/Wales/PHWC-49E443/2020|EPI_ISL_743002|2020 ···················· ···················· ···················· ···················· ···················· ····················
Z 1581 hCoV-19/Wales/PHWC-49E452/2020|EPI_ISL_743003|2020 ···················· ···················· ···················· ···················· ···················· ····················
Z 1582 hCoV-19/Wales/PHWC-49E470/2020|EPI_ISL_743005|2020 ···················· ······A············· ···················· ···················· ···················· ····················
Z 1583 hCoV-19/Wales/PHWC-49E4AD/2020|EPI_ISL_743007|2020 ···················· ···················· ···················· ···················· ···················· ····················
Z 1584 hCoV-19/Wales/PHWC-49E4BC/2020|EPI_ISL_743008|2020 ···················· ···················· ···················· ···················· ···················· ····················
Z 1585 hCoV-19/Wales/PHWC-49E4CB/2020|EPI_ISL_743009|2020 ···················· ···················· ···················· ···················· ···················· ····················
Z 1586 hCoV-19/Wales/PHWC-49E4E9/2020|EPI_ISL_743010|2020 ···················· ···················· ···················· ···················· ···················· ····················
Z 1587 hCoV-19/Wales/PHWC-49E540/2020|EPI_ISL_743013|2020 ···················· ···················· ···················· ···················· ···················· ····················
Z 1588 hCoV-19/Wales/PHWC-49E59B/2020|EPI_ISL_743016|2020 ···················· ···················· ···················· ···················· ···················· ····················
Z 1589 hCoV-19/Wales/PHWC-49E5AA/2020|EPI_ISL_743017|2020 ···················· ···················· ···················· ···················· ···················· ····················
Z 1590 hCoV-19/Wales/PHWC-49E5B9/2020|EPI_ISL_743018|2020 ···················· ···················· ···················· ···················· ···················· ····················
Z 1591 hCoV-19/Wales/PHWC-49E5C8/2020|EPI_ISL_743019|2020 ···················· ···················· ···················· ···················· ···················· ····················
Z 1592 hCoV-19/Wales/PHWC-49E5D7/2020|EPI_ISL_743020|2020 ···················· ···················· ···················· ···················· ···················· ····················
Z 1594 hCoV-19/Wales/PHWC-49E5F5/2020|EPI_ISL_743022|2020 ···················· ···················· ···················· ···················· ···················· ····················
Z 1595 hCoV-19/Wales/PHWC-49E601/2020|EPI_ISL_743023|2020 ···················· ···················· ···················· ···················· ···················· ····················
Z 1596 hCoV-19/Wales/PHWC-49E610/2020|EPI_ISL_743024|2020 ···················· ···················· ···················· ···················· ···················· ····················
Z 1597 hCoV-19/Wales/PHWC-49E62F/2020|EPI_ISL_743025|2020 ···················· ···················· ···R················ ···················· ···················· ····················
Z 1598 hCoV-19/Wales/PHWC-49E64D/2020|EPI_ISL_743027|2020 ···················· ···················· ···················· ···················· ···················· ····················
Z 1600 hCoV-19/Wales/PHWC-49E66B/2020|EPI_ISL_743029|2020 ···················· ···················· ···················· ···················· ···················· ····················
Z 1601 hCoV-19/Wales/PHWC-49E67A/2020|EPI_ISL_743030|2020 ···················· ···················· ···················· ···················· ···················· ····················
Z 1602 hCoV-19/Wales/PHWC-49E689/2020|EPI_ISL_743031|2020 ···················· ···················· ···················· ···················· ···················· ····················
Z 1603 hCoV-19/Wales/PHWC-49E698/2020|EPI_ISL_743032|2020 ···················· ···················· ···················· ···················· ···················· ····················
Z 1605 hCoV-19/Wales/PHWC-49E6D4/2020|EPI_ISL_743035|2020 ···················· ···················· ···················· ···················· ···················· ····················
Z 1606 hCoV-19/Wales/PHWC-49E6E3/2020|EPI_ISL_743036|2020 ···················· ···················· ···················· ···················· ···················· ····················
Z 1607 hCoV-19/Wales/PHWC-49E72C/2020|EPI_ISL_743039|2020 ···················· ···················· ···················· ···················· ···················· ····················
Z 1608 hCoV-19/Wales/PHWC-49E73B/2020|EPI_ISL_743040|2020 ···················· ···················· ···················· ···················· ···················· ····················
Z 1609 hCoV-19/Wales/PHWC-49E74A/2020|EPI_ISL_743041|2020 ···················· ···················· ···················· ···················· ···················· ····················
Z 1610 hCoV-19/Wales/PHWC-49E759/2020|EPI_ISL_743042|2020 ···················· ···················· ···················· ···················· ···················· ····················
Z 1611 hCoV-19/Wales/PHWC-49E768/2020|EPI_ISL_743043|2020 ···················· ···················· ···················· ···················· ···················· ····················
Z 1612 hCoV-19/Wales/PHWC-49E777/2020|EPI_ISL_743044|2020 ···················· ···················· ···················· ···················· ···················· ····················
Z 1613 hCoV-19/Wales/PHWC-49E786/2020|EPI_ISL_743045|2020 ···················· ···················· ···················· ···················· ···················· ····················
Z 1614 hCoV-19/Wales/PHWC-49E795/2020|EPI_ISL_743046|2020 ···················· ···················· ···················· ···················· ···················· ····················
Z 1615 hCoV-19/Wales/PHWC-49E7A4/2020|EPI_ISL_743047|2020 ···················· ···················· ···················· ···················· ···················· ····················
Z 1616 hCoV-19/Wales/PHWC-49E7B3/2020|EPI_ISL_743048|2020 ···················· ···················· ···················· ···················· ···················· ····················
Z 1617 hCoV-19/Wales/PHWC-49E7FF/2020|EPI_ISL_743051|2020 ···················· ···················· ···················· ···················· ···················· ····················
Z 1618 hCoV-19/Wales/PHWC-49E81A/2020|EPI_ISL_743053|2020 ···················· ···················· ···················· ···················· ···················· ····················
Z 1619 hCoV-19/Wales/PHWC-49E829/2020|EPI_ISL_743054|2020 ···················· ···················· ···················· ···················· ···················· ····················
Z 1620 hCoV-19/Wales/PHWC-49E838/2020|EPI_ISL_743055|2020 ···················· ···················· ···················· ···················· ···················· ····················
Z 1621 hCoV-19/Wales/PHWC-49E865/2020|EPI_ISL_743058|2020 ···················· ···················· ···················· ···················· ···················· ····················
Z 1622 hCoV-19/Wales/PHWC-49EB5D/2020|EPI_ISL_743099|2020 ···················· ···················· ···················· ···················· ···················· ····················
Z 1623 hCoV-19/Luxembourg/LNS5773322/2020|EPI_ISL_744300| ···················· ···················· ···················· ···················· ···················· ····················
Z 1624 hCoV-19/Luxembourg/LNS5785841/2020|EPI_ISL_744301| ···················· ···················· ···················· ···················· ···················· ····················
Z 1625 hCoV-19/Luxembourg/LNS5787107/2020|EPI_ISL_744302| ···················· ···················· ···················· ···················· ···················· ····················
Z 1626 hCoV-19/Luxembourg/LNS5800178/2020|EPI_ISL_744304| ···················· ···················· ···················· ···················· ···················· ····················
Z 1627 hCoV-19/Luxembourg/LNS5809547/2020|EPI_ISL_744305| ···················· ···················· ···················· ···················· ···················· ····················
Z 1628 hCoV-19/Luxembourg/LNS5812925/2020|EPI_ISL_744306| ···················· ···················· ···················· ···················· ···················· ····················
Z 1629 hCoV-19/Luxembourg/LNS5831514/2020|EPI_ISL_744308| ···················· ···················· ···················· ···················· ···················· ····················
Z 1630 hCoV-19/Luxembourg/LNS5855591/2020|EPI_ISL_744309| ···················· ···················· ···················· ···················· ···················· ····················
Z 1631 hCoV-19/Luxembourg/LNS5857830/2020|EPI_ISL_744310| ···················· ···················· ···················· ···················· ···················· ····················
Z 1632 hCoV-19/Luxembourg/LNS5868219/2020|EPI_ISL_744311| ···················· ···················· ···················· ···················· ···················· ····················
Z 1633 hCoV-19/Luxembourg/LNS5873219/2020|EPI_ISL_744312| ···················· ···················· ···················· ···················· ···················· ····················
Z 1634 hCoV-19/Luxembourg/LNS5882150/2020|EPI_ISL_744313| ···················· ···················· ···················· ···················· ···················· ····················
Z 1635 hCoV-19/Luxembourg/LNS5885231/2020|EPI_ISL_744314| ···················· ···················· ···················· ···················· ···················· ····················
Z 1636 hCoV-19/Luxembourg/LNS5897398/2020|EPI_ISL_744316| ···················· ···················· ···················· ···················· ······T············· ····················
Z 1637 hCoV-19/Luxembourg/LNS5897890/2020|EPI_ISL_744317| ···················· ····A··············· ···················· ···················· ···················· ····················
Z 1638 hCoV-19/Luxembourg/LNS5902641/2020|EPI_ISL_744318| ···················· ···················· ···················· ···················· ···················· ····················
Z 1639 hCoV-19/Luxembourg/LNS5905543/2020|EPI_ISL_744319| ···················· ···················· ···················· ···················· ···················· ····················
Z 1640 hCoV-19/Luxembourg/LNS5906467/2020|EPI_ISL_744320| ···················· ···················· ···················· ···················· ···················· ····················
Z 1641 hCoV-19/Luxembourg/LNS5908909/2020|EPI_ISL_744321| ···················· ···················· ···················· ···················· ···················· ····················
Z 1642 hCoV-19/Luxembourg/LNS5912684/2020|EPI_ISL_744322| ···················· G··················· ···················· ···················· ···················· ····················
Z 1643 hCoV-19/Luxembourg/LNS5913589/2020|EPI_ISL_744323| ···················· ···················· ···················· ···················· ···················· ····················
Z 1644 hCoV-19/Luxembourg/LNS5921376/2020|EPI_ISL_744324| ···················· ···················· ···················· ···················· ···················· ····················
Z 1645 hCoV-19/Luxembourg/LNS5927877/2020|EPI_ISL_744325| ···················· ···················· ···················· ···················· ······T············· ····················
Z 1646 hCoV-19/Luxembourg/LNS5936010/2020|EPI_ISL_744326| ···················· ···················· ···················· ···················· ···················· ····················
Z 1647 hCoV-19/Luxembourg/LNS5936689/2020|EPI_ISL_744327| ···················· ···················· ···················· ···················· ···················· ····················
Z 1648 hCoV-19/Luxembourg/LNS5948760/2020|EPI_ISL_744328| ···················· G··················· ···················· ···················· ···················· ····················
Z 1649 hCoV-19/Luxembourg/LNS5966835/2020|EPI_ISL_744331| ···················· ···················· ···················· ···················· ···················· ····················
```

| | | | | | | | | |
|---|---|---|---|---|---|---|---|---|
| Z | 1650 | hCoV-19/Luxembourg/LNS5980660/2020\|EPI_ISL_744332\| | ···················· | ···················· | ···················· | ···················· | ···················· | ···················· |
| Z | 1651 | hCoV-19/Luxembourg/LNS5986534/2020\|EPI_ISL_744333\| | ···················· | ····A··············· | ···················· | ···················· | ···················· | ···················· |
| Z | 1652 | hCoV-19/Luxembourg/LNS5988821/2020\|EPI_ISL_744334\| | ···················· | ···················· | ···················· | ···················· | ···················· | ···················· |
| Z | 1653 | hCoV-19/Luxembourg/LNS5994653/2020\|EPI_ISL_744335\| | ···················· | ···················· | ···················· | ···················· | ···················· | ···················· |
| Z | 1654 | hCoV-19/Luxembourg/LNS6015230/2020\|EPI_ISL_744337\| | ···················· | ···················· | ···················· | ···················· | ······T············· | ···················· |
| Z | 1655 | hCoV-19/Luxembourg/LNS6023868/2020\|EPI_ISL_744338\| | ···················· | ···················· | ···················· | ···················· | ···················· | ···················· |
| Z | 1656 | hCoV-19/Luxembourg/LNS6027069/2020\|EPI_ISL_744339\| | ···················· | ···················· | ···················· | ···················· | ···················· | ···················· |
| Z | 1657 | hCoV-19/Luxembourg/LNS6028262/2020\|EPI_ISL_744340\| | ···················· | ···················· | ···················· | ···················· | ···················· | ···················· |
| Z | 1658 | hCoV-19/Luxembourg/LNS6032919/2020\|EPI_ISL_744341\| | ···················· | ···················· | ···················· | ···················· | ···················· | ···················· |
| Z | 1659 | hCoV-19/Luxembourg/LNS6034314/2020\|EPI_ISL_744342\| | ···················· | ···················· | ···················· | ···················· | ···················· | ···················· |
| Z | 1660 | hCoV-19/Luxembourg/LNS6043596/2020\|EPI_ISL_744343\| | ···················· | ···················· | ···················· | ···················· | ···················· | ···················· |
| Z | 1661 | hCoV-19/Luxembourg/LNS6044069/2020\|EPI_ISL_744344\| | ···················· | ···················· | ···················· | ···················· | ···················· | ···················· |
| Z | 1662 | hCoV-19/Luxembourg/LNS6046845/2020\|EPI_ISL_744345\| | ···················· | ···················· | ···················· | ···················· | ···················· | ···················· |
| Z | 1663 | hCoV-19/Luxembourg/LNS6050650/2020\|EPI_ISL_744346\| | ···················· | ···················· | ···················· | ···················· | ···················· | ···················· |
| Z | 1664 | hCoV-19/Luxembourg/LNS6058776/2020\|EPI_ISL_744347\| | ···················· | ···················· | ···················· | ···················· | ···················· | ···················· |
| Z | 1665 | hCoV-19/Luxembourg/LNS6067075/2020\|EPI_ISL_744349\| | ···················· | ···················· | ···················· | ···················· | ···················· | ···················· |
| Z | 1666 | hCoV-19/Luxembourg/LNS6071962/2020\|EPI_ISL_744350\| | ···················· | ···················· | ···················· | ···················· | ···················· | ···················· |
| Z | 1667 | hCoV-19/Luxembourg/LNS6073102/2020\|EPI_ISL_744351\| | ···················· | ···················· | ···················· | ···················· | ···················· | ···················· |
| Z | 1668 | hCoV-19/Luxembourg/LNS6079656/2020\|EPI_ISL_744352\| | ···················· | ···················· | ···················· | ···················· | ···················· | ···················· |
| Z | 1669 | hCoV-19/Luxembourg/LNS6079739/2020\|EPI_ISL_744353\| | ···················· | ···················· | ···················· | ···················· | ···················· | ···················· |
| Z | 1670 | hCoV-19/Luxembourg/LNS6079967/2020\|EPI_ISL_744354\| | ···················· | ···················· | ···················· | ···················· | ···················· | ···················· |
| Z | 1671 | hCoV-19/Luxembourg/LNS6089147/2020\|EPI_ISL_744355\| | ···················· | ···················· | ···················· | ···················· | ···················· | ···················· |
| Z | 1672 | hCoV-19/Luxembourg/LNS6089486/2020\|EPI_ISL_744356\| | ···················· | ···················· | ···················· | ···················· | ···················· | ···················· |
| Z | 1673 | hCoV-19/Luxembourg/LNS6090235/2020\|EPI_ISL_744357\| | ···················· | ···················· | ···················· | ···················· | ···················· | ···················· |
| Z | 1674 | hCoV-19/Luxembourg/LNS6092237/2020\|EPI_ISL_744358\| | ···················· | ···················· | ···················· | ···················· | ···················· | ···················· |
| Z | 1675 | hCoV-19/Luxembourg/LNS6092797/2020\|EPI_ISL_744359\| | ···················· | ···················· | ···················· | ···················· | ···················· | ···················· |
| Z | 1676 | hCoV-19/Luxembourg/LNS6106493/2020\|EPI_ISL_744360\| | ···················· | ···················· | ···················· | ···················· | ···················· | ···················· |
| Z | 1677 | hCoV-19/Luxembourg/LNS6109598/2020\|EPI_ISL_744361\| | ···················· | ···················· | ···················· | ···················· | ···················· | ···················· |
| Z | 1678 | hCoV-19/Luxembourg/LNS6113575/2020\|EPI_ISL_744362\| | ···················· | ···················· | ···················· | ···················· | ···················· | ···················· |
| Z | 1679 | hCoV-19/Luxembourg/LNS6115650/2020\|EPI_ISL_744363\| | ···················· | ···················· | ···················· | ···················· | ···················· | ···················· |
| Z | 1680 | hCoV-19/Luxembourg/LNS6115978/2020\|EPI_ISL_744364\| | ···················· | ···················· | ···················· | ···················· | ···················· | ···················· |
| Z | 1681 | hCoV-19/Luxembourg/LNS6117100/2020\|EPI_ISL_744365\| | ···················· | ···················· | ···················· | ···················· | ···················· | ···················· |
| Z | 1682 | hCoV-19/Luxembourg/LNS6120480/2020\|EPI_ISL_744366\| | ···················· | G··················· | ···················· | ···················· | ···················· | ···················· |
| Z | 1683 | hCoV-19/Luxembourg/LNS6133647/2020\|EPI_ISL_744367\| | ···················· | ···················· | ···················· | ···················· | ···················· | ···················· |
| Z | 1684 | hCoV-19/Luxembourg/LNS6137065/2020\|EPI_ISL_744369\| | ···················· | ···················· | ···················· | ···················· | ···················· | ···················· |
| Z | 1685 | hCoV-19/Luxembourg/LNS6164249/2020\|EPI_ISL_744370\| | ···················· | ···················· | ···················· | ···················· | ···················· | ···················· |
| Z | 1686 | hCoV-19/Luxembourg/LNS6166486/2020\|EPI_ISL_744372\| | ···················· | ···················· | ···················· | ···················· | ···················· | ···················· |
| Z | 1687 | hCoV-19/Luxembourg/LNS6172864/2020\|EPI_ISL_744373\| | ···················· | ···················· | ···················· | ···················· | ···················· | ···················· |
| Z | 1688 | hCoV-19/Luxembourg/LNS6173898/2020\|EPI_ISL_744374\| | ···················· | ···················· | ···················· | ···················· | ···················· | ···················· |
| Z | 1689 | hCoV-19/Luxembourg/LNS6175625/2020\|EPI_ISL_744375\| | ···················· | ···················· | ···················· | ···················· | ···················· | ···················· |
| Z | 1690 | hCoV-19/Luxembourg/LNS6183746/2020\|EPI_ISL_744376\| | ···················· | ···················· | ···················· | ···················· | ···················· | ···················· |
| Z | 1691 | hCoV-19/Luxembourg/LNS6186776/2020\|EPI_ISL_744378\| | ···················· | ···················· | ···················· | ···················· | ···················· | ···················· |
| Z | 1692 | hCoV-19/Luxembourg/LNS6199114/2020\|EPI_ISL_744379\| | ···················· | ···················· | ···················· | ···················· | ···················· | ···················· |
| Z | 1693 | hCoV-19/Luxembourg/LNS6199628/2020\|EPI_ISL_744380\| | ···················· | ···················· | ···················· | ···················· | ···················· | ···················· |
| Z | 1694 | hCoV-19/Luxembourg/LNS6200431/2020\|EPI_ISL_744381\| | ···················· | ···················· | ···················· | ···················· | ···················· | ···················· |
| Z | 1695 | hCoV-19/Luxembourg/LNS6216319/2020\|EPI_ISL_744383\| | ···················· | ···················· | ···················· | ···················· | ···················· | ···················· |
| Z | 1696 | hCoV-19/Luxembourg/LNS6229527/2020\|EPI_ISL_744385\| | ···················· | ···················· | ···················· | ···················· | ···················· | ···················· |
| Z | 1697 | hCoV-19/Luxembourg/LNS6230945/2020\|EPI_ISL_744386\| | ···················· | ···················· | ···················· | ···················· | ···················· | ···················· |
| Z | 1698 | hCoV-19/Luxembourg/LNS6235819/2020\|EPI_ISL_744387\| | ···················· | ···················· | ···················· | ···················· | ···················· | ···················· |
| Z | 1699 | hCoV-19/Luxembourg/LNS6237904/2020\|EPI_ISL_744388\| | ···················· | ···················· | ···················· | ···················· | ······T············· | ···················· |
| Z | 1700 | hCoV-19/Luxembourg/LNS6246495/2020\|EPI_ISL_744389\| | ···················· | ···················· | ···················· | ···················· | ···················· | ···················· |
| Z | 1701 | hCoV-19/USA/UT-UPHL-2012332988/2020\|EPI_ISL_745659 | ···················· | ···················· | ···················· | ···················· | ···················· | ···················· |
| Z | 1702 | hCoV-19/USA/UT-UPHL-2012827437/2020\|EPI_ISL_745660 | ···················· | ·A·················· | ···················· | ···················· | ···················· | ···················· |
| Z | 1703 | hCoV-19/USA/UT-UPHL-2012396895/2020\|EPI_ISL_745661 | ···················· | ···················· | ···················· | ···················· | ···················· | ···················· |
| Z | 1704 | hCoV-19/USA/UT-UPHL-2012170875/2020\|EPI_ISL_745662 | ···················· | ···················· | ···················· | ···················· | ···················· | ···················· |
| Z | 1705 | hCoV-19/USA/UT-UPHL-2012692663/2020\|EPI_ISL_745663 | ···················· | ···················· | ···················· | ···················· | ···················· | ···················· |
| Z | 1706 | hCoV-19/USA/UT-UPHL-2012728359/2020\|EPI_ISL_745664 | ···················· | ···················· | ···················· | ···················· | ···················· | ···················· |
| Z | 1707 | hCoV-19/USA/UT-UPHL-2012159735/2020\|EPI_ISL_745665 | ···················· | ···················· | ···················· | ···················· | ···················· | ···················· |
| Z | 1708 | hCoV-19/USA/UT-UPHL-2012579514/2020\|EPI_ISL_745666 | ···················· | ···················· | ···················· | ···················· | ···················· | ···················· |
| Z | 1709 | hCoV-19/USA/UT-UPHL-2012855927/2020\|EPI_ISL_745667 | ···················· | ···················· | ···················· | ···················· | ···················· | ···················· |
| Z | 1710 | hCoV-19/USA/UT-UPHL-2012831200/2020\|EPI_ISL_745668 | ···················· | ···················· | ···················· | ···················· | ···················· | ···················· |
| Z | 1711 | hCoV-19/USA/UT-UPHL-2012291444/2020\|EPI_ISL_745669 | ···················· | ···················· | ···················· | ···················· | ···················· | ···················· |
| Z | 1712 | hCoV-19/USA/UT-UPHL-2012097796/2020\|EPI_ISL_745671 | ···················· | ···················· | ···················· | ···················· | ···················· | ···················· |
| Z | 1713 | hCoV-19/USA/UT-UPHL-2012530976/2020\|EPI_ISL_745672 | ···················· | ···················· | ···················· | ···················· | ···················· | ···················· |
| Z | 1714 | hCoV-19/USA/UT-UPHL-2012590314/2020\|EPI_ISL_745673 | ···················· | ···················· | ···················· | ···················· | ···················· | ···················· |
| Z | 1715 | hCoV-19/USA/UT-UPHL-2012096893/2020\|EPI_ISL_745674 | ···················· | ·A·················· | ···················· | ···················· | ···················· | ···················· |
| Z | 1716 | hCoV-19/USA/UT-UPHL-2012401784/2020\|EPI_ISL_745675 | ···················· | ···················· | ···················· | ···················· | ···················· | ···················· |
| Z | 1717 | hCoV-19/USA/UT-UPHL-2012069277/2020\|EPI_ISL_745676 | ···················· | ···················· | ···················· | ···················· | ···················· | ···················· |
| Z | 1718 | hCoV-19/USA/UT-UPHL-2012647991/2020\|EPI_ISL_745677 | ···················· | ···················· | ···················· | ···················· | ···················· | ···················· |
| Z | 1719 | hCoV-19/USA/UT-UPHL-2012126681/2020\|EPI_ISL_745678 | ···················· | ·A·················· | ···················· | ···················· | ···················· | ···················· |
| Z | 1720 | hCoV-19/USA/UT-UPHL-2012449428/2020\|EPI_ISL_745679 | ···················· | ···················· | ···················· | ···················· | ···················· | ···················· |
| Z | 1721 | hCoV-19/USA/UT-UPHL-2012475809/2020\|EPI_ISL_745680 | ···················· | ···················· | ···················· | ···················· | ···················· | ···················· |
| Z | 1722 | hCoV-19/USA/UT-UPHL-2012408644/2020\|EPI_ISL_745681 | ···················· | ···················· | ···················· | ···················· | ···················· | ···················· |
| Z | 1723 | hCoV-19/USA/UT-UPHL-2012568306/2020\|EPI_ISL_745682 | ···················· | ···················· | ···················· | ···················· | ···················· | ···················· |
| Z | 1724 | hCoV-19/USA/UT-UPHL-2012562063/2020\|EPI_ISL_745683 | ···················· | ·A·················· | ···················· | ···················· | ···················· | ···················· |
| Z | 1725 | hCoV-19/USA/UT-UPHL-2012385708/2020\|EPI_ISL_745684 | ···················· | ···················· | ···················· | ···················· | ···················· | ···················· |
| Z | 1726 | hCoV-19/USA/UT-UPHL-2012990874/2020\|EPI_ISL_745685 | ···················· | ···················· | ···················· | ···················· | ···················· | ···················· |
| Z | 1727 | hCoV-19/USA/UT-UPHL-2012756486/2020\|EPI_ISL_745686 | ···················· | ···················· | ···················· | ···················· | ···················· | ···················· |
| Z | 1728 | hCoV-19/USA/UT-UPHL-2012152848/2020\|EPI_ISL_745687 | ···················· | ···················· | ···················· | ···················· | ···················· | ···················· |
| Z | 1729 | hCoV-19/USA/UT-UPHL-2012142641/2020\|EPI_ISL_745688 | ···················· | ···················· | ···················· | ···················· | ···················· | ···················· |

| | | | | | | | | |
|---|---|---|---|---|---|---|---|---|
| Z | 1730 | hCoV-19/USA/UT-UPHL-2012519731/2020\|EPI_ISL_745689 | ···················· | ·A·················· | ····················· | ···················· | ···················· | ···················· |
| Z | 1731 | hCoV-19/USA/UT-UPHL-2012885396/2020\|EPI_ISL_745690 | ···················· | ···················· | ····················· | ···················· | ···················· | ···················· |
| Z | 1732 | hCoV-19/USA/UT-UPHL-2012140285/2020\|EPI_ISL_745691 | ···················· | ···················· | ····················· | ···················· | ···················· | ···················· |
| Z | 1733 | hCoV-19/USA/UT-UPHL-2012330509/2020\|EPI_ISL_745692 | ···················· | ···················· | ····················· | ···················· | ···················· | ···················· |
| Z | 1734 | hCoV-19/USA/UT-UPHL-2012807270/2020\|EPI_ISL_745693 | ···················· | ···················· | ····················· | ···················· | ···················· | ···················· |
| Z | 1735 | hCoV-19/USA/UT-UPHL-2012066229/2020\|EPI_ISL_745694 | ···················· | ···················· | ····················· | ···················· | ···················· | ···················· |
| Z | 1736 | hCoV-19/USA/UT-UPHL-2012227413/2020\|EPI_ISL_745695 | ···················· | ···················· | ····················· | ···················· | ···················· | ···················· |
| Z | 1737 | hCoV-19/USA/UT-UPHL-2012145786/2020\|EPI_ISL_745696 | ···················· | ···················· | ····················· | ···················· | ···················· | ···················· |
| Z | 1738 | hCoV-19/USA/UT-UPHL-2012609427/2020\|EPI_ISL_745697 | ···················· | ·A·················· | ····················· | ···················· | ···················· | ···················· |
| Z | 1739 | hCoV-19/USA/UT-UPHL-2012140722/2020\|EPI_ISL_745698 | ···················· | ·A·················· | ····················· | ···················· | ···················· | ···················· |
| Z | 1740 | hCoV-19/USA/UT-UPHL-2012040948/2020\|EPI_ISL_745699 | ···················· | ···················· | ····················· | ···················· | ···················· | ···················· |
| Z | 1741 | hCoV-19/USA/UT-UPHL-2012821012/2020\|EPI_ISL_746948 | ···················· | ·A·················· | ····················· | ···················· | ···················· | ···················· |
| Z | 1742 | hCoV-19/USA/UT-UPHL-2012804386/2020\|EPI_ISL_746949 | ···················· | ·A·················· | ····················· | ···················· | ···················· | ···················· |
| Z | 1743 | hCoV-19/USA/UT-UPHL-2012522594/2020\|EPI_ISL_746950 | ···················· | ···················· | ····················· | ···················· | ···················· | ···················· |
| Z | 1744 | hCoV-19/USA/UT-UPHL-2012537908/2020\|EPI_ISL_746951 | ···················· | ·A·················· | ····················· | ···················· | ···················· | ···················· |
| Z | 1745 | hCoV-19/USA/UT-UPHL-2012116193/2020\|EPI_ISL_746952 | ···················· | ·A·················· | ····················· | ···················· | ···················· | ···················· |
| Z | 1746 | hCoV-19/USA/UT-UPHL-2012819565/2020\|EPI_ISL_746953 | ···················· | ·A·················· | ····················· | ···················· | ···················· | ···················· |
| Z | 1747 | hCoV-19/USA/UT-UPHL-2012913150/2020\|EPI_ISL_746954 | ···················· | ·A·················· | ····················· | ···················· | ···················· | ···················· |
| Z | 1748 | hCoV-19/USA/UT-UPHL-2012089416/2020\|EPI_ISL_746955 | ···················· | ·A·················· | ····················· | ···················· | ···················· | ···················· |
| Z | 1749 | hCoV-19/USA/UT-UPHL-2012172138/2020\|EPI_ISL_746956 | ···················· | ·A·················· | ····················· | ···················· | ···················· | ···················· |
| Z | 1750 | hCoV-19/USA/UT-UPHL-2012414402/2020\|EPI_ISL_746957 | ···················· | ·A·················· | ····················· | ···················· | ···················· | ···················· |
| Z | 1751 | hCoV-19/USA/UT-UPHL-2012453307/2020\|EPI_ISL_746958 | ···················· | ···················· | ····················· | ···················· | ···················· | ···················· |
| Z | 1752 | hCoV-19/USA/UT-UPHL-2012057649/2020\|EPI_ISL_746959 | ···················· | ···················· | ····················· | ···················· | ···················· | ···················· |
| Z | 1753 | hCoV-19/USA/UT-UPHL-2012189554/2020\|EPI_ISL_746960 | ···················· | ·A·················· | ····················· | ···················· | ···················· | ···················· |
| Z | 1754 | hCoV-19/USA/UT-UPHL-2012119230/2020\|EPI_ISL_746961 | ···················· | ·A·················· | ····················· | ···················· | ···················· | ···················· |
| Z | 1755 | hCoV-19/USA/UT-UPHL-2012146142/2020\|EPI_ISL_746962 | ···················· | ·A·················· | ····················· | ···················· | ···················· | ···················· |
| Z | 1756 | hCoV-19/USA/UT-UPHL-2012463625/2020\|EPI_ISL_746963 | ···················· | ···················· | ····················· | ···················· | ···················· | ···················· |
| Z | 1757 | hCoV-19/USA/UT-UPHL-2012301287/2020\|EPI_ISL_746964 | ···················· | ·A·················· | ····················· | ···················· | ···················· | ···················· |
| Z | 1758 | hCoV-19/USA/UT-UPHL-2012893348/2020\|EPI_ISL_746965 | ···················· | ·A·················· | ····················· | ···················· | ···················· | ···················· |
| Z | 1759 | hCoV-19/USA/UT-UPHL-2012248421/2020\|EPI_ISL_746966 | ···················· | ···················· | ····················· | ···················· | ···················· | ···················· |
| Z | 1760 | hCoV-19/USA/UT-UPHL-2012650580/2020\|EPI_ISL_746967 | ···················· | ···················· | ····················· | ···················· | ···················· | ···················· |
| Z | 1761 | hCoV-19/USA/UT-UPHL-2012720932/2020\|EPI_ISL_746968 | ···················· | ···················· | ····················· | ···················· | ···················· | ···················· |
| Z | 1762 | hCoV-19/USA/UT-UPHL-2012392301/2020\|EPI_ISL_746969 | ···················· | ···················· | ····················· | ···················· | ···················· | ···················· |
| Z | 1763 | hCoV-19/USA/UT-UPHL-2012273960/2020\|EPI_ISL_746970 | ···················· | ·A·················· | ····················· | ···················· | ···················· | ···················· |
| Z | 1764 | hCoV-19/USA/UT-UPHL-2012530254/2020\|EPI_ISL_746971 | ···················· | ···················· | ····················· | ···················· | ···················· | ···················· |
| Z | 1765 | hCoV-19/USA/UT-UPHL-2012355224/2020\|EPI_ISL_746972 | ···················· | ···················· | ····················· | ···················· | ···················· | ···················· |
| Z | 1766 | hCoV-19/USA/UT-UPHL-2012811501/2020\|EPI_ISL_746973 | ···················· | ···················· | ····················· | ···················· | ···················· | ···················· |
| Z | 1767 | hCoV-19/USA/UT-UPHL-2012618844/2020\|EPI_ISL_746974 | ···················· | ·A·················· | ····················· | ···················· | ···················· | ···················· |
| Z | 1768 | hCoV-19/USA/UT-UPHL-2012124802/2020\|EPI_ISL_746975 | ···················· | ···················· | ····················· | ···················· | ···················· | ···················· |
| Z | 1769 | hCoV-19/USA/UT-UPHL-2012152791/2020\|EPI_ISL_746976 | ···················· | ·A·················· | ····················· | ···················· | ···················· | ···················· |
| Z | 1770 | hCoV-19/USA/UT-UPHL-2012642336/2020\|EPI_ISL_746977 | ···················· | ···················· | ····················· | ···················· | ···················· | ···················· |
| Z | 1771 | hCoV-19/USA/UT-UPHL-2012474685/2020\|EPI_ISL_746978 | ···················· | ···················· | ····················· | ···················· | ···················· | ···················· |
| Z | 1772 | hCoV-19/USA/UT-UPHL-2012123020/2020\|EPI_ISL_746979 | ···················· | ·A·················· | ····················· | ···················· | ···················· | ···················· |
| Z | 1773 | hCoV-19/USA/UT-UPHL-2012784062/2020\|EPI_ISL_746980 | ···················· | ·A·················· | ····················· | ···················· | ···················· | ···················· |
| Z | 1774 | hCoV-19/USA/UT-UPHL-2012694228/2020\|EPI_ISL_746981 | ···················· | ···················· | ····················· | ···················· | ···················· | ···················· |
| Z | 1775 | hCoV-19/USA/UT-UPHL-2012775940/2020\|EPI_ISL_746982 | ···················· | ·A·················· | ····················· | ···················· | ···················· | ···················· |
| Z | 1776 | hCoV-19/USA/UT-UPHL-2012254967/2020\|EPI_ISL_746983 | ···················· | ·A·················· | ····················· | ···················· | ···················· | ···················· |
| Z | 1777 | hCoV-19/USA/UT-UPHL-2012261094/2020\|EPI_ISL_746984 | ···················· | ···················· | ····················· | ···················· | ···················· | ···················· |
| Z | 1778 | hCoV-19/USA/UT-UPHL-2012116492/2020\|EPI_ISL_746985 | ···················· | ···················· | ····················· | ···················· | ···················· | ···················· |
| Z | 1779 | hCoV-19/USA/UT-UPHL-2012453686/2020\|EPI_ISL_746986 | ···················· | ···················· | ····················· | ···················· | ···················· | ···················· |
| Z | 1780 | hCoV-19/USA/UT-UPHL-2012421387/2020\|EPI_ISL_746987 | ···················· | ···················· | ····················· | ···················· | ···················· | ···················· |
| Z | 1781 | hCoV-19/USA/UT-UPHL-2012275347/2020\|EPI_ISL_746988 | ···················· | ···················· | ····················· | ···················· | ···················· | ···················· |
| Z | 1782 | hCoV-19/USA/UT-UPHL-2012982319/2020\|EPI_ISL_746989 | ···················· | ···················· | ····················· | ···················· | ···················· | ···················· |
| Z | 1783 | hCoV-19/USA/UT-UPHL-2012685593/2020\|EPI_ISL_746990 | ···················· | ···················· | ····················· | ···················· | ···················· | ···················· |
| Z | 1784 | hCoV-19/USA/UT-UPHL-2012413942/2020\|EPI_ISL_746991 | ···················· | ·A·················· | ····················· | ···················· | ···················· | ···················· |
| Z | 1785 | hCoV-19/USA/UT-UPHL-2012922673/2020\|EPI_ISL_746992 | ···················· | ·A·················· | ····················· | ···················· | ···················· | ···················· |
| Z | 1786 | hCoV-19/USA/UT-UPHL-2012802557/2020\|EPI_ISL_746993 | ···················· | ···················· | ····················· | ···················· | ···················· | ···················· |
| Z | 1787 | hCoV-19/USA/UT-UPHL-2012376464/2020\|EPI_ISL_746994 | ···················· | ·A·················· | ····················· | ···················· | ···················· | ···················· |
| Z | 1788 | hCoV-19/USA/UT-UPHL-2012338760/2020\|EPI_ISL_746995 | ···················· | ···················· | ····················· | ···················· | ···················· | ···················· |
| Z | 1789 | hCoV-19/USA/UT-UPHL-2012209911/2020\|EPI_ISL_746996 | ···················· | ···················· | ····················· | ···················· | ···················· | ···················· |
| Z | 1790 | hCoV-19/USA/UT-UPHL-2012265204/2020\|EPI_ISL_746997 | ···················· | ·A·················· | ····················· | ···················· | ···················· | ···················· |
| Z | 1791 | hCoV-19/USA/UT-UPHL-2012104383/2020\|EPI_ISL_746998 | ···················· | ···················· | ····················· | ···················· | ···················· | ···················· |
| Z | 1792 | hCoV-19/USA/UT-UPHL-2012533316/2020\|EPI_ISL_746999 | ···················· | ···················· | ····················· | ···················· | ···················· | ···················· |
| Z | 1793 | hCoV-19/Luxembourg/LNS0568422/2020\|EPI_ISL_739800\| | ···················· | ···················· | ····················· | ···················· | ···················· | ···················· |
| Z | 1794 | hCoV-19/Luxembourg/LNS0587977/2020\|EPI_ISL_739801\| | ···················· | ···················· | ····················· | ···················· | ···················· | ···················· |
| Z | 1795 | hCoV-19/Luxembourg/LNS0589025/2020\|EPI_ISL_739802\| | ···················· | ···················· | ····················· | ···················· | ······T············· | ···················· |
| Z | 1796 | hCoV-19/Luxembourg/LNS0600176/2020\|EPI_ISL_739803\| | ···················· | ···················· | ····················· | ···················· | ···················· | ···················· |
| Z | 1797 | hCoV-19/Luxembourg/LNS0601879/2020\|EPI_ISL_739804\| | ···················· | ···················· | ····················· | ···················· | ···················· | ···················· |
| Z | 1798 | hCoV-19/Luxembourg/LNS0613111/2020\|EPI_ISL_739805\| | ···················· | ···················· | ····················· | ···················· | ···················· | ···················· |
| Z | 1799 | hCoV-19/Luxembourg/LNS0617896/2020\|EPI_ISL_739806\| | ···················· | ···················· | ····················· | ···················· | ···················· | ···················· |
| Z | 1800 | hCoV-19/Luxembourg/LNS0623813/2020\|EPI_ISL_739807\| | ···················· | ···················· | ····················· | ···················· | ···················· | ···················· |
| Z | 1801 | hCoV-19/Luxembourg/LNS0642518/2020\|EPI_ISL_739809\| | ···················· | ···················· | ····················· | ···················· | ···················· | ···················· |
| Z | 1802 | hCoV-19/Luxembourg/LNS0654077/2020\|EPI_ISL_739810\| | ···················· | ···················· | ····················· | ···················· | ···················· | ···················· |
| Z | 1803 | hCoV-19/Luxembourg/LNS0655598/2020\|EPI_ISL_739811\| | ···················· | G··················· | ····················· | ···················· | ···················· | ···················· |
| Z | 1804 | hCoV-19/Luxembourg/LNS0655930/2020\|EPI_ISL_739812\| | ···················· | ···················· | ····················· | ···················· | ······T············· | ···················· |
| Z | 1805 | hCoV-19/Luxembourg/LNS0673237/2020\|EPI_ISL_739814\| | ···················· | ···················· | ····················· | ···················· | ···················· | ···················· |
| Z | 1806 | hCoV-19/Luxembourg/LNS0674406/2020\|EPI_ISL_739815\| | ···················· | ···················· | ····················· | ···················· | ···················· | ···················· |
| Z | 1807 | hCoV-19/Luxembourg/LNS0682941/2020\|EPI_ISL_739816\| | ···················· | ···················· | ····················· | ···················· | ···················· | ···················· |
| Z | 1808 | hCoV-19/Luxembourg/LNS0694152/2020\|EPI_ISL_739817\| | ···················· | ···················· | ····················· | ···················· | ···················· | ···················· |
| Z | 1809 | hCoV-19/Luxembourg/LNS0694160/2020\|EPI_ISL_739818\| | ···················· | ···················· | ····················· | ···················· | ···················· | ···················· |

| | | | | | | | | |
|---|---|---|---|---|---|---|---|---|
| Z | 1810 | hCoV-19/Luxembourg/LNS0700306/2020\|EPI_ISL_739819\| | ···················· | ···················· | ···················· | ···················· | ···················· | ···················· |
| Z | 1811 | hCoV-19/Luxembourg/LNS0721959/2020\|EPI_ISL_739820\| | ···················· | ···················· | ···················· | ···················· | ···················· | ···················· |
| Z | 1812 | hCoV-19/Luxembourg/LNS0725139/2020\|EPI_ISL_739821\| | ···················· | ···················· | ···················· | ···················· | ···················· | ···················· |
| Z | 1813 | hCoV-19/Luxembourg/LNS0726804/2020\|EPI_ISL_739822\| | ···················· | ···················· | ···················· | ···················· | ···················· | ···················· |
| Z | 1814 | hCoV-19/Luxembourg/LNS0730142/2020\|EPI_ISL_739823\| | ···················· | ···················· | ···················· | ···················· | ···················· | ···················· |
| Z | 1815 | hCoV-19/Luxembourg/LNS0734464/2020\|EPI_ISL_739824\| | ···················· | ···················· | ···················· | ···················· | ···················· | ···················· |
| Z | 1816 | hCoV-19/Luxembourg/LNS0747135/2020\|EPI_ISL_739825\| | ···················· | ···················· | ···················· | ···················· | ···················· | ···················· |
| Z | 1817 | hCoV-19/Luxembourg/LNS0751416/2020\|EPI_ISL_739826\| | ···················· | ·A·················· | ···················· | ···················· | ···················· | ···················· |
| Z | 1818 | hCoV-19/Luxembourg/LNS0754234/2020\|EPI_ISL_739827\| | ···················· | ···················· | ···················· | ···················· | ···················· | ···················· |
| Z | 1819 | hCoV-19/Luxembourg/LNS0756714/2020\|EPI_ISL_739828\| | ···················· | ···················· | ···················· | ···················· | ···················· | ···················· |
| Z | 1820 | hCoV-19/Luxembourg/LNS0759351/2020\|EPI_ISL_739829\| | ···················· | ···················· | ···················· | ···················· | ···················· | ···················· |
| Z | 1821 | hCoV-19/Luxembourg/LNS0772976/2020\|EPI_ISL_739830\| | ···················· | ···················· | ···················· | ···················· | ·····T·············· | ···················· |
| Z | 1822 | hCoV-19/Luxembourg/LNS0779841/2020\|EPI_ISL_739831\| | ···················· | ···················· | ···················· | ···················· | ···················· | ···················· |
| Z | 1823 | hCoV-19/Luxembourg/LNS0795813/2020\|EPI_ISL_739832\| | ···················· | ···················· | ···················· | ···················· | ···················· | ···················· |
| Z | 1824 | hCoV-19/Luxembourg/LNS0797488/2020\|EPI_ISL_739833\| | ···················· | ···················· | ···················· | ···················· | ···················· | ···················· |
| Z | 1825 | hCoV-19/Luxembourg/LNS0801829/2020\|EPI_ISL_739834\| | ···················· | ···················· | ···················· | ···················· | ···················· | ···················· |
| Z | 1826 | hCoV-19/Luxembourg/LNS0815952/2020\|EPI_ISL_739835\| | ···················· | ···················· | ···················· | ···················· | ···················· | ···················· |
| Z | 1827 | hCoV-19/Luxembourg/LNS0829711/2020\|EPI_ISL_739836\| | ···················· | ···················· | ···················· | ···················· | ·····T·············· | ···················· |
| Z | 1828 | hCoV-19/Luxembourg/LNS0831693/2020\|EPI_ISL_739837\| | ···················· | ···················· | ···················· | ···················· | ···················· | ···················· |
| Z | 1829 | hCoV-19/Luxembourg/LNS0842029/2020\|EPI_ISL_739839\| | ···················· | ···················· | ···················· | ···················· | ···················· | ···················· |
| Z | 1830 | hCoV-19/Luxembourg/LNS0843817/2020\|EPI_ISL_739840\| | ···················· | ···················· | ···················· | ···················· | ···················· | ···················· |
| Z | 1831 | hCoV-19/Luxembourg/LNS0849119/2020\|EPI_ISL_739841\| | ···················· | ···················· | ···················· | ···················· | ···················· | ···················· |
| Z | 1832 | hCoV-19/Luxembourg/LNS0853178/2020\|EPI_ISL_739842\| | ···················· | ···················· | ···················· | ···················· | ···················· | ···················· |
| Z | 1833 | hCoV-19/Luxembourg/LNS0876336/2020\|EPI_ISL_739844\| | ···················· | ···················· | ···················· | ···················· | ···················· | ···················· |
| Z | 1834 | hCoV-19/Luxembourg/LNS0884586/2020\|EPI_ISL_739845\| | ···················· | ···················· | ···················· | ···················· | ···················· | ···················· |
| Z | 1835 | hCoV-19/Luxembourg/LNS0889216/2020\|EPI_ISL_739846\| | ···················· | ···················· | ···················· | ···················· | ···················· | ···················· |
| Z | 1836 | hCoV-19/Luxembourg/LNS0893820/2020\|EPI_ISL_739847\| | ···················· | ···················· | ···················· | ···················· | ···················· | ···················· |
| Z | 1837 | hCoV-19/Luxembourg/LNS0895040/2020\|EPI_ISL_739848\| | ···················· | ···················· | ···················· | ···················· | ···················· | ···················· |
| Z | 1838 | hCoV-19/Luxembourg/LNS0901454/2020\|EPI_ISL_739849\| | ···················· | ···················· | ···················· | ···················· | ···················· | ···················· |
| Z | 1839 | hCoV-19/Luxembourg/LNS0903249/2020\|EPI_ISL_739850\| | ···················· | ···················· | ···················· | ···················· | ···················· | ···················· |
| Z | 1840 | hCoV-19/Luxembourg/LNS0908586/2020\|EPI_ISL_739851\| | ···················· | ···················· | ···················· | ···················· | ···················· | ···················· |
| Z | 1841 | hCoV-19/Luxembourg/LNS0910783/2020\|EPI_ISL_739852\| | ···················· | ···················· | ···················· | ···················· | ···················· | ···················· |
| Z | 1842 | hCoV-19/Luxembourg/LNS0912617/2020\|EPI_ISL_739853\| | ···················· | ···················· | ···················· | ···················· | ···················· | ···················· |
| Z | 1843 | hCoV-19/Luxembourg/LNS0913789/2020\|EPI_ISL_739854\| | ···················· | ···················· | ···················· | ···················· | ···················· | ···················· |
| Z | 1844 | hCoV-19/Luxembourg/LNS0914893/2020\|EPI_ISL_739855\| | ···················· | ···················· | ···················· | ···················· | ···················· | ···················· |
| Z | 1845 | hCoV-19/Luxembourg/LNS0917978/2020\|EPI_ISL_739856\| | ···················· | ···················· | ···················· | ···················· | ···················· | ···················· |
| Z | 1846 | hCoV-19/Luxembourg/LNS0921251/2020\|EPI_ISL_739857\| | ···················· | ···················· | ···················· | ···················· | ···················· | ···················· |
| Z | 1847 | hCoV-19/Luxembourg/LNS0927824/2020\|EPI_ISL_739858\| | ···················· | ···················· | ···················· | ···················· | ···················· | ···················· |
| Z | 1848 | hCoV-19/Luxembourg/LNS0928223/2020\|EPI_ISL_739859\| | ···················· | ···A················ | ···················· | ···················· | ···················· | ···················· |
| Z | 1849 | hCoV-19/Luxembourg/LNS0936084/2020\|EPI_ISL_739860\| | ···················· | ···················· | ···················· | ···················· | ···················· | ···················· |
| Z | 1850 | hCoV-19/Luxembourg/LNS0947533/2020\|EPI_ISL_739861\| | ···················· | ···················· | ···················· | ···················· | ···················· | ···················· |
| Z | 1851 | hCoV-19/Luxembourg/LNS0958161/2020\|EPI_ISL_739862\| | ···················· | ···················· | ···················· | ···················· | ···················· | ···················· |
| Z | 1852 | hCoV-19/Luxembourg/LNS0965116/2020\|EPI_ISL_739863\| | ···················· | ···················· | ···················· | ···················· | ···················· | ···················· |
| Z | 1853 | hCoV-19/Luxembourg/LNS0968104/2020\|EPI_ISL_739864\| | ···················· | ···················· | ···················· | ···················· | ···················· | ···················· |
| Z | 1854 | hCoV-19/Luxembourg/LNS0971128/2020\|EPI_ISL_739865\| | ···················· | ···················· | ···················· | ···················· | ···················· | ···················· |
| Z | 1855 | hCoV-19/Luxembourg/LNS0985896/2020\|EPI_ISL_739867\| | ···················· | ···················· | ···················· | ···················· | ···················· | ···················· |
| Z | 1856 | hCoV-19/Luxembourg/LNS0988203/2020\|EPI_ISL_739868\| | ···················· | ···················· | ···················· | ···················· | ···················· | ···················· |
| Z | 1857 | hCoV-19/Luxembourg/LNS0994663/2020\|EPI_ISL_739869\| | ···················· | ···················· | ···················· | ···················· | ···················· | ···················· |
| Z | 1858 | hCoV-19/Luxembourg/LNS1014689/2020\|EPI_ISL_739870\| | ···················· | ···················· | ···················· | ···················· | ···················· | ···················· |
| Z | 1859 | hCoV-19/Luxembourg/LNS1019791/2020\|EPI_ISL_739871\| | ···················· | ···················· | ···················· | ···················· | ···················· | ···················· |
| Z | 1860 | hCoV-19/Luxembourg/LNS1021995/2020\|EPI_ISL_739872\| | ···················· | G··················· | ···················· | ···················· | ···················· | ···················· |
| Z | 1861 | hCoV-19/Luxembourg/LNS1024274/2020\|EPI_ISL_739873\| | ···················· | ···················· | ···················· | ···················· | ···················· | ···················· |
| Z | 1862 | hCoV-19/Luxembourg/LNS1028885/2020\|EPI_ISL_739874\| | ···················· | ···················· | ···················· | ···················· | ···················· | ···················· |
| Z | 1863 | hCoV-19/Luxembourg/LNS1030661/2020\|EPI_ISL_739875\| | ···················· | ···················· | ···················· | ···················· | ···················· | ···················· |
| Z | 1864 | hCoV-19/Luxembourg/LNS1034986/2020\|EPI_ISL_739876\| | ···················· | ···················· | ···················· | ···················· | ···················· | ···················· |
| Z | 1865 | hCoV-19/Luxembourg/LNS1044750/2020\|EPI_ISL_739878\| | ···················· | ···················· | ···················· | ···················· | ···················· | ···················· |
| Z | 1866 | hCoV-19/Luxembourg/LNS1049978/2020\|EPI_ISL_739879\| | ···················· | ···················· | ···················· | ···················· | ···················· | ···················· |
| Z | 1867 | hCoV-19/Luxembourg/LNS1050653/2020\|EPI_ISL_739881\| | ···················· | ···················· | ···················· | ···················· | ···················· | ···················· |
| Z | 1868 | hCoV-19/Luxembourg/LNS1057454/2020\|EPI_ISL_739882\| | ···················· | ···················· | ···················· | ···················· | ···················· | ···················· |
| Z | 1869 | hCoV-19/Luxembourg/LNS1059363/2020\|EPI_ISL_739883\| | ···················· | ···················· | ···················· | ···················· | ···················· | ···················· |
| Z | 1870 | hCoV-19/Luxembourg/LNS1060278/2020\|EPI_ISL_739884\| | ···················· | ···················· | ···················· | ···················· | ···················· | ···················· |
| Z | 1871 | hCoV-19/Luxembourg/LNS1078153/2020\|EPI_ISL_739885\| | ···················· | ···················· | ···················· | ···················· | ···················· | ···················· |
| Z | 1872 | hCoV-19/Luxembourg/LNS1096019/2020\|EPI_ISL_739886\| | ···················· | ···················· | ···················· | ···················· | ·····T·············· | ···················· |
| Z | 1873 | hCoV-19/Luxembourg/LNS1102691/2020\|EPI_ISL_739887\| | ···················· | ···················· | ···················· | ···················· | ···················· | ···················· |
| Z | 1874 | hCoV-19/Luxembourg/LNS1110082/2020\|EPI_ISL_739888\| | ···················· | ···················· | ···················· | ···················· | ···················· | ···················· |
| Z | 1875 | hCoV-19/Luxembourg/LNS1111678/2020\|EPI_ISL_739889\| | ···················· | ···················· | ···················· | ···················· | ·····T·············· | ···················· |
| Z | 1876 | hCoV-19/Luxembourg/LNS1113037/2020\|EPI_ISL_739890\| | ···················· | ···A················ | ···················· | ···················· | ···················· | ···················· |
| Z | 1877 | hCoV-19/Luxembourg/LNS1118090/2020\|EPI_ISL_739891\| | ···················· | ···················· | ···················· | ···················· | ···················· | ···················· |
| Z | 1878 | hCoV-19/Luxembourg/LNS1124969/2020\|EPI_ISL_739892\| | ···················· | ···················· | ···················· | ···················· | ···················· | ···················· |
| Z | 1879 | hCoV-19/Luxembourg/LNS1128482/2020\|EPI_ISL_739893\| | ···················· | ···················· | ···················· | ···················· | ···················· | ···················· |
| Z | 1880 | hCoV-19/Luxembourg/LNS1131770/2020\|EPI_ISL_739894\| | ···················· | ···················· | ···················· | ···················· | ···················· | ···················· |
| Z | 1881 | hCoV-19/Luxembourg/LNS1146047/2020\|EPI_ISL_739896\| | ···················· | ···················· | ···················· | ···················· | ···················· | ···················· |
| Z | 1882 | hCoV-19/Luxembourg/LNS1148230/2020\|EPI_ISL_739897\| | ···················· | ···················· | ···················· | ···················· | ···················· | ···················· |
| Z | 1883 | hCoV-19/Luxembourg/LNS1149202/2020\|EPI_ISL_739898\| | ···················· | ···················· | ···················· | ···················· | ···················· | ···················· |
| Z | 1884 | hCoV-19/Luxembourg/LNS1152992/2020\|EPI_ISL_739899\| | ···················· | ···················· | ···················· | ···················· | ···················· | ···················· |
| Z | 1885 | hCoV-19/Canada/BC-BCCDC-8435/2020\|EPI_ISL_740800\|2 | ···················· | ···················· | ···················· | ···················· | ···················· | ···················· |
| Z | 1886 | hCoV-19/Canada/BC-BCCDC-8437/2020\|EPI_ISL_740801\|2 | ···················· | ···················· | ···················· | ···················· | ···················· | ···················· |
| Z | 1887 | hCoV-19/Canada/BC-BCCDC-8440/2020\|EPI_ISL_740802\|2 | ···················· | ···················· | ···················· | ···················· | ···················· | ···················· |
| Z | 1888 | hCoV-19/Canada/BC-BCCDC-8441/2020\|EPI_ISL_740803\|2 | ···················· | ···················· | ···················· | ···················· | ···················· | ···················· |
| Z | 1889 | hCoV-19/Canada/BC-BCCDC-8443/2020\|EPI_ISL_740804\|2 | ···················· | ···················· | ···················· | ···················· | ···················· | ···················· |

| | | | | | | | |
|---|---|---|---|---|---|---|---|
| Z | 1890 hCoV-19/Canada/BC-BCCDC-8444/2020\|EPI_ISL_740805\|2 | ···················· | ·A·················· | ···················· | ···················· | ···················· | ··A················· |
| Z | 1891 hCoV-19/Canada/BC-BCCDC-8447/2020\|EPI_ISL_740806\|2 | ···················· | ···················· | ···················· | ···················· | ···················· | ···················· |
| Z | 1892 hCoV-19/Canada/BC-BCCDC-8449/2020\|EPI_ISL_740807\|2 | ···················· | ···················· | ···················· | ···················· | ···················· | ···················· |
| Z | 1893 hCoV-19/Canada/BC-BCCDC-8544/2020\|EPI_ISL_740808\|2 | ···················· | ···················· | ···················· | ···················· | ···················· | ···················· |
| Z | 1894 hCoV-19/Canada/BC-BCCDC-8545/2020\|EPI_ISL_740809\|2 | ···················· | ···················· | ···················· | ···················· | ···················· | ···················· |
| Z | 1895 hCoV-19/Canada/BC-BCCDC-8546/2020\|EPI_ISL_740810\|2 | ···················· | ···················· | ···················· | ···················· | ···················· | ···················· |
| Z | 1896 hCoV-19/Canada/BC-BCCDC-8547/2020\|EPI_ISL_740811\|2 | ···················· | ···················· | ···················· | ···················· | ···················· | ···················· |
| Z | 1897 hCoV-19/Canada/BC-BCCDC-8548/2020\|EPI_ISL_740812\|2 | ···················· | ···················· | ···················· | ···················· | ···················· | ···················· |
| Z | 1898 hCoV-19/Canada/BC-BCCDC-8549/2020\|EPI_ISL_740813\|2 | ···················· | ···················· | ···················· | ···················· | ···················· | ···················· |
| Z | 1899 hCoV-19/Canada/BC-BCCDC-8550/2020\|EPI_ISL_740814\|2 | ···················· | ·A·················· | ···················· | ···················· | ···················· | ··A················· |
| Z | 1900 hCoV-19/Canada/BC-BCCDC-8555/2020\|EPI_ISL_740815\|2 | ···················· | ···················· | ···················· | ···················· | ···················· | ···················· |
| Z | 1901 hCoV-19/Canada/BC-BCCDC-8557/2020\|EPI_ISL_740816\|2 | ···················· | ···················· | ···················· | ···················· | ···················· | ···················· |
| Z | 1902 hCoV-19/Canada/BC-BCCDC-8558/2020\|EPI_ISL_740817\|2 | ···················· | ···················· | ···················· | ···················· | ···················· | ···················· |
| Z | 1903 hCoV-19/Canada/BC-BCCDC-8559/2020\|EPI_ISL_740818\|2 | ···················· | ···················· | ···················· | ···················· | ···················· | ···················· |
| Z | 1904 hCoV-19/Canada/BC-BCCDC-8560/2020\|EPI_ISL_740819\|2 | ···················· | ···················· | ···················· | ···················· | ···················· | ···················· |
| Z | 1905 hCoV-19/Canada/BC-BCCDC-8561/2020\|EPI_ISL_740820\|2 | ···················· | ···················· | ···················· | ···················· | ···················· | ···················· |
| Z | 1906 hCoV-19/Canada/BC-BCCDC-8562/2020\|EPI_ISL_740821\|2 | ···················· | ·A·················· | ···················· | ···················· | ···················· | ··A················· |
| Z | 1907 hCoV-19/Canada/BC-BCCDC-8563/2020\|EPI_ISL_740822\|2 | ···················· | ···················· | ···················· | ···················· | ···················· | ···················· |
| Z | 1908 hCoV-19/Canada/BC-BCCDC-8564/2020\|EPI_ISL_740823\|2 | ···················· | ·A·················· | ···················· | ···················· | ···················· | ··A················· |
| Z | 1909 hCoV-19/Canada/BC-BCCDC-8565/2020\|EPI_ISL_740824\|2 | ···················· | ···················· | ···················· | ···················· | ···················· | ···················· |
| Z | 1910 hCoV-19/Canada/BC-BCCDC-8566/2020\|EPI_ISL_740825\|2 | ···················· | ·A·················· | ···················· | ···················· | ···················· | ··A················· |
| Z | 1911 hCoV-19/Canada/BC-BCCDC-8576/2020\|EPI_ISL_740826\|2 | ···················· | ·A·················· | ···················· | ···················· | ···················· | ··A················· |
| Z | 1912 hCoV-19/Canada/BC-BCCDC-8577/2020\|EPI_ISL_740827\|2 | ···················· | ·A·················· | ···················· | ···················· | ···················· | ··A················· |
| Z | 1913 hCoV-19/Canada/BC-BCCDC-8578/2020\|EPI_ISL_740828\|2 | ···················· | ·A·················· | ···················· | ···················· | ···················· | ··A················· |
| Z | 1914 hCoV-19/Canada/BC-BCCDC-8579/2020\|EPI_ISL_740829\|2 | ···················· | ·A·················· | ···················· | ···················· | ···················· | ··A················· |
| Z | 1915 hCoV-19/Canada/BC-BCCDC-8581/2020\|EPI_ISL_740830\|2 | ···················· | ·A·················· | ···················· | ···················· | ···················· | ··A················· |
| Z | 1916 hCoV-19/Canada/BC-BCCDC-8584/2020\|EPI_ISL_740831\|2 | ···················· | ·A·················· | ···················· | ···················· | ···················· | ··A················· |
| Z | 1917 hCoV-19/Canada/BC-BCCDC-8586/2020\|EPI_ISL_740833\|2 | ···················· | ···················· | ···················· | ···················· | ···················· | ···················· |
| Z | 1918 hCoV-19/Canada/BC-BCCDC-8587/2020\|EPI_ISL_740834\|2 | ···················· | ·A·················· | ···················· | ···················· | ···················· | ··A················· |
| Z | 1919 hCoV-19/Canada/BC-BCCDC-8588/2020\|EPI_ISL_740835\|2 | ···················· | ···················· | ···················· | ···················· | ···················· | ···················· |
| Z | 1920 hCoV-19/Canada/BC-BCCDC-8589/2020\|EPI_ISL_740836\|2 | ···················· | ···················· | ···················· | ···················· | ···················· | ···················· |
| Z | 1921 hCoV-19/Canada/BC-BCCDC-8590/2020\|EPI_ISL_740837\|2 | ···················· | ···················· | ···················· | ···················· | ···················· | ···················· |
| Z | 1922 hCoV-19/Canada/BC-BCCDC-8591/2020\|EPI_ISL_740838\|2 | ···················· | ···················· | ···················· | ···················· | ···················· | ···················· |
| Z | 1923 hCoV-19/Canada/BC-BCCDC-8593/2020\|EPI_ISL_740840\|2 | ···················· | ···················· | ···················· | ···················· | ···················· | ···················· |
| Z | 1924 hCoV-19/Canada/BC-BCCDC-8595/2020\|EPI_ISL_740841\|2 | ···················· | ···················· | ···················· | ···················· | ···················· | ···················· |
| Z | 1925 hCoV-19/Canada/BC-BCCDC-8597/2020\|EPI_ISL_740842\|2 | ···················· | ···················· | ···················· | ···················· | ···················· | ···················· |
| Z | 1926 hCoV-19/Canada/BC-BCCDC-8598/2020\|EPI_ISL_740843\|2 | ···················· | ···················· | ···················· | ···················· | ···················· | ···················· |
| Z | 1927 hCoV-19/Canada/BC-BCCDC-8600/2020\|EPI_ISL_740844\|2 | ···················· | ···················· | ···················· | ···················· | ···················· | ···················· |
| Z | 1928 hCoV-19/Canada/BC-BCCDC-8601/2020\|EPI_ISL_740845\|2 | ···················· | ·A·················· | ···················· | ···················· | ···················· | ··A················· |
| Z | 1929 hCoV-19/Canada/BC-BCCDC-8603/2020\|EPI_ISL_740846\|2 | ···················· | ···················· | ···················· | ···················· | ···················· | ···················· |
| Z | 1930 hCoV-19/Canada/BC-BCCDC-8604/2020\|EPI_ISL_740847\|2 | ···················· | ···················· | ···················· | ···················· | ···················· | ···················· |
| Z | 1931 hCoV-19/Canada/BC-BCCDC-8607/2020\|EPI_ISL_740849\|2 | ···················· | ···················· | ···················· | ···················· | ···················· | ···················· |
| Z | 1932 hCoV-19/Canada/BC-BCCDC-8608/2020\|EPI_ISL_740850\|2 | ···················· | ·A·················· | ···················· | ···················· | ···················· | ··A················· |
| Z | 1933 hCoV-19/Canada/BC-BCCDC-8612/2020\|EPI_ISL_740851\|2 | ···················· | ···················· | ···················· | ···················· | ···················· | ···················· |
| Z | 1934 hCoV-19/Canada/BC-BCCDC-8616/2020\|EPI_ISL_740853\|2 | ···················· | ·A·················· | ···················· | ···················· | ···················· | ··A················· |
| Z | 1935 hCoV-19/Canada/BC-BCCDC-8619/2020\|EPI_ISL_740855\|2 | ···················· | ·A·················· | ···················· | ···················· | ···················· | ··A················· |
| Z | 1936 hCoV-19/Canada/BC-BCCDC-8621/2020\|EPI_ISL_740856\|2 | ···················· | ···················· | ···················· | ···················· | ···················· | ···················· |
| Z | 1937 hCoV-19/Canada/BC-BCCDC-8625/2020\|EPI_ISL_740857\|2 | ···················· | ·A·················· | ···················· | ···················· | ···················· | ··A················· |
| Z | 1938 hCoV-19/Canada/BC-BCCDC-8627/2020\|EPI_ISL_740858\|2 | ···················· | ···················· | ···················· | ···················· | ···················· | ···················· |
| Z | 1939 hCoV-19/Canada/BC-BCCDC-8633/2020\|EPI_ISL_740859\|2 | ···················· | ···················· | ···················· | ···················· | ···················· | ···················· |
| Z | 1940 hCoV-19/Canada/BC-BCCDC-8635/2020\|EPI_ISL_740860\|2 | ···················· | ···················· | ···················· | ···················· | ···················· | ···················· |
| Z | 1941 hCoV-19/Canada/BC-BCCDC-8636/2020\|EPI_ISL_740861\|2 | ···················· | ···················· | ···················· | ···················· | ···················· | ···················· |
| Z | 1942 hCoV-19/Canada/BC-BCCDC-8637/2020\|EPI_ISL_740862\|2 | ···················· | ···················· | ···················· | ···················· | ···················· | ···················· |
| Z | 1943 hCoV-19/Australia/NSW1232/2020\|EPI_ISL_740863\|2020 | ···················· | ···················· | ···················· | ···················· | ···················· | ···················· |
| Z | 1944 hCoV-19/Australia/NSW1233/2020\|EPI_ISL_740864\|2020 | ···················· | ···················· | ···················· | ···················· | ···················· | ···················· |
| Z | 1945 hCoV-19/Australia/NSW1235/2020\|EPI_ISL_740866\|2020 | ···················· | ···················· | ···················· | ···················· | ···················· | ···················· |
| Z | 1946 hCoV-19/Australia/NSW1236/2020\|EPI_ISL_740867\|2020 | ···················· | ···················· | ···················· | ···················· | ···················· | ···················· |
| Z | 1947 hCoV-19/Australia/NSW1237/2020\|EPI_ISL_740868\|2020 | ···················· | ·········A·········· | ···················· | ···················· | ···················· | ···················· |
| Z | 1948 hCoV-19/Australia/NSW1238/2020\|EPI_ISL_740869\|2020 | ···················· | ···················· | ···················· | ···················· | ···················· | ···················· |
| Z | 1949 hCoV-19/Australia/NSW1239/2020\|EPI_ISL_740870\|2020 | ···················· | ···················· | ···················· | ···················· | ···················· | ···················· |
| Z | 1950 hCoV-19/Australia/NSW1240/2020\|EPI_ISL_740871\|2020 | ···················· | ···················· | ···················· | ···················· | ···················· | ···················· |
| Z | 1951 hCoV-19/Australia/NSW1241/2020\|EPI_ISL_740872\|2020 | ···················· | ···················· | ···················· | ···················· | ···················· | ···················· |
| Z | 1952 hCoV-19/Australia/NSW1243/2020\|EPI_ISL_740874\|2020 | ···················· | ·A·················· | ···················· | ···················· | ···················· | ··A················· |
| Z | 1953 hCoV-19/Australia/NSW1244/2020\|EPI_ISL_740875\|2020 | ···················· | ···················· | ···················· | ···················· | ···················· | ···················· |
| Z | 1954 hCoV-19/Australia/NSW1245/2020\|EPI_ISL_740876\|2020 | ···················· | ···················· | ···················· | ···················· | ···················· | ···················· |
| Z | 1955 hCoV-19/Australia/NSW1246/2020\|EPI_ISL_740877\|2020 | ···················· | ···················· | ···················· | ···················· | ···················· | ···················· |
| Z | 1956 hCoV-19/Australia/NSW1247/2020\|EPI_ISL_740878\|2020 | ···················· | ···················· | ···················· | ···················· | ···················· | ···················· |
| Z | 1957 hCoV-19/Australia/NSW1248/2020\|EPI_ISL_740879\|2020 | ···················· | ···················· | ···················· | ···················· | ···················· | ···················· |
| Z | 1958 hCoV-19/Australia/NSW1249/2020\|EPI_ISL_740880\|2020 | ···················· | ···················· | ···················· | ···················· | ···················· | ···················· |
| Z | 1959 hCoV-19/Australia/NSW1250/2020\|EPI_ISL_740881\|2020 | ···················· | ···················· | ···················· | ···················· | ···················· | ···················· |
| Z | 1960 hCoV-19/Australia/NSW1251/2020\|EPI_ISL_740882\|2020 | ···················· | ···················· | ···················· | ···················· | ···················· | ···················· |
| Z | 1961 hCoV-19/Australia/NSW1252/2020\|EPI_ISL_740883\|2020 | ···················· | ···················· | ···················· | ···················· | ···················· | ···················· |
| Z | 1962 hCoV-19/Australia/NSW1253/2020\|EPI_ISL_740884\|2020 | ···················· | ···················· | ···················· | ···················· | ···················· | ···················· |
| Z | 1963 hCoV-19/Australia/NSW1254/2020\|EPI_ISL_740885\|2020 | ···················· | ···················· | ···················· | ···················· | ···················· | ···················· |
| Z | 1964 hCoV-19/Australia/NSW1255/2020\|EPI_ISL_740886\|2020 | ···················· | ···················· | ···················· | ···················· | ···················· | ···················· |
| Z | 1965 hCoV-19/Australia/NSW1256/2020\|EPI_ISL_740887\|2020 | ···················· | ···················· | ···················· | ···················· | ···················· | ···················· |
| Z | 1966 hCoV-19/Australia/NSW1257/2020\|EPI_ISL_740888\|2020 | ···················· | ···················· | ···················· | ···················· | ···················· | ···················· |
| Z | 1967 hCoV-19/Australia/NSW1258/2020\|EPI_ISL_740889\|2020 | ···················· | ···················· | ···················· | ···················· | ···················· | ···················· |
| Z | 1968 hCoV-19/Australia/NSW1259/2020\|EPI_ISL_740890\|2020 | ···················· | ···················· | ···················· | ···················· | ···················· | ···················· |
| Z | 1969 hCoV-19/Australia/NSW1260/2020\|EPI_ISL_740891\|2020 | ···················· | ···················· | ···················· | ···················· | ···················· | ···················· |

Z 1970 hCoV-19/Australia/NSW1261/2020|EPI_ISL_740892|2020 ···················· ···················· ···················· ···················· ···················· ····················
Z 1971 hCoV-19/Australia/NSW1262/2020|EPI_ISL_740893|2020 ···················· ···················· ···················· ···················· ···················· ····················
Z 1972 hCoV-19/Australia/NSW1263/2020|EPI_ISL_740894|2020 ···················· ···················· ···················· ···················· ···················· ····················
Z 1973 hCoV-19/Australia/NSW1264/2020|EPI_ISL_740895|2020 ···················· ···················· ···················· ···················· ···················· ····················
Z 1974 hCoV-19/Australia/NSW1265/2020|EPI_ISL_740896|2020 ···················· ···················· ···················· ···················· ···················· ····················
Z 1975 hCoV-19/England/ALDP-B750FC/2020|EPI_ISL_740897|20 ···················· ···················· ···················· ···················· ···················· ····················
Z 1976 hCoV-19/England/ALDP-B75108/2020|EPI_ISL_740898|20 ···················· ···················· ···················· ···················· ···················· ····················
Z 1977 hCoV-19/England/ALDP-B751EA/2020|EPI_ISL_740899|20 ···················· ···················· ···················· ···················· ···················· ····················
Z 1978 hCoV-19/Wales/PHWC-49E892/2020|EPI_ISL_743060|2020 ···················· ···················· ···················· ···················· ···················· ····················
Z 1979 hCoV-19/Wales/PHWC-49E8A1/2020|EPI_ISL_743061|2020 ···················· ···················· ···················· ···················· ···················· ····················
Z 1980 hCoV-19/Wales/PHWC-49E8DE/2020|EPI_ISL_743063|2020 ···················· ···················· ···················· ···················· ···················· ····················
Z 1981 hCoV-19/Wales/PHWC-49E8ED/2020|EPI_ISL_743064|2020 ···················· ···················· ···················· ···················· ···················· ····················
Z 1982 hCoV-19/Wales/PHWC-49E8FC/2020|EPI_ISL_743065|2020 ···················· ···················· ···················· ···················· ···················· ····················
Z 1983 hCoV-19/Wales/PHWC-49E908/2020|EPI_ISL_743066|2020 ···················· ···················· ···················· ···················· ···················· ···A················
Z 1984 hCoV-19/Wales/PHWC-49E917/2020|EPI_ISL_743067|2020 ···················· ···················· ···················· ···················· ···················· ····················
Z 1985 hCoV-19/Wales/PHWC-49E926/2020|EPI_ISL_743068|2020 ···················· ···················· ···················· ···················· ···················· ····················
Z 1986 hCoV-19/Wales/PHWC-49E944/2020|EPI_ISL_743070|2020 ···················· ···················· ···················· ···················· ···················· ····················
Z 1987 hCoV-19/Wales/PHWC-49E962/2020|EPI_ISL_743072|2020 ···················· ···················· ···················· ···················· ···················· ····················
Z 1988 hCoV-19/Wales/PHWC-49E971/2020|EPI_ISL_743073|2020 ···················· ···················· ···················· ···················· ···················· ····················
Z 1989 hCoV-19/Wales/PHWC-49E9AE/2020|EPI_ISL_743076|2020 ···················· ···················· ···················· ···················· ···················· ····················
Z 1990 hCoV-19/Wales/PHWC-49E9CC/2020|EPI_ISL_743078|2020 ···················· ···················· ···················· ···················· ···················· ····················
Z 1991 hCoV-19/Wales/PHWC-49E9F9/2020|EPI_ISL_743080|2020 ···················· ···················· ···················· ···················· ···················· ····················
Z 1992 hCoV-19/Wales/PHWC-49EA14/2020|EPI_ISL_743082|2020 ···················· ···················· ···················· ···················· ···················· ···A················
Z 1993 hCoV-19/Wales/PHWC-49EA32/2020|EPI_ISL_743084|2020 ···················· ···················· ···················· ···················· ···················· ····················
Z 1994 hCoV-19/Wales/PHWC-49EA41/2020|EPI_ISL_743085|2020 ·T·················· ···················· ···················· ···················· ···················· ····················
Z 1995 hCoV-19/Wales/PHWC-49EA8D/2020|EPI_ISL_743089|2020 ···················· ···················· ···················· ···················· ···················· ····················
Z 1996 hCoV-19/Wales/PHWC-49EABA/2020|EPI_ISL_743091|2020 ···················· ···················· ···················· ···················· ···················· ····················
Z 1997 hCoV-19/Wales/PHWC-49EAC9/2020|EPI_ISL_743092|2020 ···················· ···················· ···················· ···················· ···················· ····················
Z 1998 hCoV-19/Wales/PHWC-49EAD8/2020|EPI_ISL_743093|2020 ···················· ···················· ···················· ···················· ···················· ····················
Z 1999 hCoV-19/Wales/PHWC-49EAE7/2020|EPI_ISL_743094|2020 ···················· ···················· ···················· ···················· ···················· ····················
Z 2000 hCoV-19/Wales/PHWC-49EB02/2020|EPI_ISL_743095|2020 ···················· ···················· ···················· ···················· ···················· ····················
Z 2001 hCoV-19/Wales/PHWC-49EB11/2020|EPI_ISL_743096|2020 ···················· ···················· ···················· ···················· ···················· ····················
Z 2002 hCoV-19/Wales/PHWC-49EB4E/2020|EPI_ISL_743098|2020 ···················· ···················· ···················· ···················· ···················· ····················
Z 2003 hCoV-19/Luxembourg/LNS6246850/2020|EPI_ISL_744390| ···················· ····A··············· ···················· ···················· ···················· ····················
Z 2004 hCoV-19/Luxembourg/LNS6247688/2020|EPI_ISL_744391| ···················· ···················· ···················· ···················· ···················· ····················
Z 2005 hCoV-19/Luxembourg/LNS6258184/2020|EPI_ISL_744392| ···················· ···················· ···················· ···················· ···················· ····················
Z 2006 hCoV-19/Luxembourg/LNS6262346/2020|EPI_ISL_744393| ···················· ···················· ···················· ···················· ···················· ····················
Z 2007 hCoV-19/Luxembourg/LNS6262705/2020|EPI_ISL_744394| ···················· ···················· ···················· ···················· ···················· ····················
Z 2008 hCoV-19/Luxembourg/LNS6275805/2020|EPI_ISL_744395| ···················· ···················· ···················· ···················· ···················· ····················
Z 2009 hCoV-19/Luxembourg/LNS6286268/2020|EPI_ISL_744396| ···················· ···················· ···················· ···················· ···················· ····················
Z 2010 hCoV-19/Luxembourg/LNS6322021/2020|EPI_ISL_744397| ···················· ···················· ···················· ···················· ···················· ····················
Z 2011 hCoV-19/Luxembourg/LNS6323748/2020|EPI_ISL_744398| ···················· ···················· ···················· ···················· ···················· ····················
Z 2012 hCoV-19/Luxembourg/LNS6327525/2020|EPI_ISL_744399| ···················· ···················· ···················· ···················· ···················· ····················
Z 2013 hCoV-19/Luxembourg/LNS6424507/2020|EPI_ISL_744410| ···················· ···················· ···················· ···················· ···················· ····················
Z 2014 hCoV-19/Luxembourg/LNS6449164/2020|EPI_ISL_744416| ···················· ···················· ···················· ···················· ···················· ····················
Z 2015 hCoV-19/Luxembourg/LNS6449725/2020|EPI_ISL_744417| ···················· ···················· ···················· ···················· ···················· ····················
Z 2016 hCoV-19/Luxembourg/LNS6454473/2020|EPI_ISL_744418| ···················· ···················· ···················· ···················· ···················· ····················
Z 2017 hCoV-19/Luxembourg/LNS6477547/2020|EPI_ISL_744420| ···················· ···················· ···················· ···················· ···················· ····················
Z 2018 hCoV-19/Luxembourg/LNS6510114/2020|EPI_ISL_744423| ···················· ···················· ···················· ···················· ···················· ····················
Z 2019 hCoV-19/Luxembourg/LNS6512343/2020|EPI_ISL_744424| ···················· ···················· ···················· ···················· ···················· ····················
Z 2020 hCoV-19/Luxembourg/LNS6516073/2020|EPI_ISL_744425| ···················· ···················· ···················· ···················· ···················· ····················
Z 2021 hCoV-19/Luxembourg/LNS6526267/2020|EPI_ISL_744426| ···················· ···················· ···················· ···················· ···················· ····················
Z 2022 hCoV-19/USA/UT-UPHL-2012486257/2020|EPI_ISL_745700 ···················· ···················· ···················· ···················· ···················· ····················
Z 2023 hCoV-19/USA/UT-UPHL-2012657838/2020|EPI_ISL_745701 ···················· ···················· ···················· ···················· ···················· ····················
Z 2024 hCoV-19/USA/UT-UPHL-2012689482/2020|EPI_ISL_745702 ···················· ···················· ···················· ···················· ···················· ····················
Z 2025 hCoV-19/USA/UT-UPHL-2012104720/2020|EPI_ISL_745703 ···················· ···················· ···················· ···················· ···················· ····················
Z 2026 hCoV-19/USA/UT-UPHL-2012025968/2020|EPI_ISL_745704 ···················· ···················· ···················· ···················· ···················· ····················
Z 2027 hCoV-19/USA/UT-UPHL-2012038111/2020|EPI_ISL_745705 ···················· ·A·················· ···················· ···················· ···················· ····················
Z 2028 hCoV-19/USA/UT-UPHL-2012403517/2020|EPI_ISL_745706 ···················· ···················· ···················· ···················· ···················· ····················
Z 2029 hCoV-19/USA/UT-UPHL-2012343311/2020|EPI_ISL_745707 ···················· ···················· ···················· ···················· ···················· ····················
Z 2030 hCoV-19/USA/UT-UPHL-2012769332/2020|EPI_ISL_745708 ···················· ···················· ···················· ···················· ···················· ····················
Z 2031 hCoV-19/USA/UT-UPHL-2012546162/2020|EPI_ISL_745709 ···················· ···················· ···················· ···················· ···················· ····················
Z 2032 hCoV-19/USA/UT-UPHL-2012034164/2020|EPI_ISL_745710 ···················· ···················· ···················· ···················· ···················· ····················
Z 2033 hCoV-19/USA/UT-UPHL-2012907859/2020|EPI_ISL_745711 ···················· ···················· ···················· ···················· ···················· ····················
Z 2034 hCoV-19/USA/UT-UPHL-2012125058/2020|EPI_ISL_745712 ···················· ···················· ···················· ···················· ···················· ····················
Z 2035 hCoV-19/USA/UT-UPHL-2012751236/2020|EPI_ISL_745713 ···················· ···················· ···················· ···················· ···················· ····················
Z 2036 hCoV-19/USA/UT-UPHL-2012541662/2020|EPI_ISL_745741 ···················· ···················· ···················· ···················· ···················· ····················
Z 2037 hCoV-19/USA/UT-UPHL-2012807295/2020|EPI_ISL_745742 ···················· ···················· ···················· ···················· ···················· ····················
Z 2038 hCoV-19/USA/UT-UPHL-2012529217/2020|EPI_ISL_745747 ···················· ···················· ···················· ···················· ···················· ····················
Z 2039 hCoV-19/USA/UT-UPHL-2012804969/2020|EPI_ISL_745748 ···················· ···················· ···················· ···················· ···················· ····················
Z 2040 hCoV-19/USA/UT-UPHL-2012652830/2020|EPI_ISL_745749 ···················· ···················· ···················· ···················· ···················· ····················
Z 2041 hCoV-19/USA/UT-UPHL-2012776293/2020|EPI_ISL_745750 ···················· ···················· ···················· ···················· ···················· ····················
Z 2042 hCoV-19/USA/UT-UPHL-2012523098/2020|EPI_ISL_745751 ···················· ···················· ···················· ···················· ···················· ····················
Z 2043 hCoV-19/USA/UT-UPHL-2012079566/2020|EPI_ISL_745752 ···················· ···················· ···················· ···················· ···················· ····················
Z 2044 hCoV-19/USA/UT-UPHL-2012697266/2020|EPI_ISL_745753 ···················· ···················· ···················· ···················· ···················· ····················
Z 2045 hCoV-19/USA/UT-UPHL-2012296348/2020|EPI_ISL_745754 ···················· ···················· ···················· ···················· ···················· ····················
Z 2046 hCoV-19/USA/UT-UPHL-2012050843/2020|EPI_ISL_745755 ···················· ·A·················· ···················· ···················· ···················· ····················
Z 2047 hCoV-19/USA/UT-UPHL-2012006772/2020|EPI_ISL_745756 ···················· ···················· ···················· ···················· ···················· ····················
Z 2048 hCoV-19/USA/UT-UPHL-2012437468/2020|EPI_ISL_745757 ···················· ···················· ···················· ···················· ···················· ····················
Z 2049 hCoV-19/Luxembourg/LNS1162404/2020|EPI_ISL_739901| ···················· ···················· ···················· ···················· ···················· ····················

| | | | | | | | |
|---|---|---|---|---|---|---|---|
| Z | 2050 hCoV-19/Luxembourg/LNS1162436/2020\|EPI_ISL_739902\| | ···················· | ···················· | ····················· | ···················· | ···················· | ···················· |
| Z | 2051 hCoV-19/Luxembourg/LNS1165717/2020\|EPI_ISL_739903\| | ···················· | ···················· | ····················· | ···················· | ······T············· | ···················· |
| Z | 2052 hCoV-19/Luxembourg/LNS1174515/2020\|EPI_ISL_739904\| | ···················· | ···················· | ····················· | ···················· | ···················· | ···················· |
| Z | 2053 hCoV-19/Luxembourg/LNS1186930/2020\|EPI_ISL_739905\| | ···················· | ···················· | ····················· | ···················· | ···················· | ···················· |
| Z | 2054 hCoV-19/Luxembourg/LNS1190671/2020\|EPI_ISL_739906\| | ···················· | ···················· | ····················· | ···················· | ···················· | ···················· |
| Z | 2055 hCoV-19/Luxembourg/LNS1202747/2020\|EPI_ISL_739907\| | ···················· | ···················· | ····················· | ···················· | ···················· | ···················· |
| Z | 2056 hCoV-19/Luxembourg/LNS1203891/2020\|EPI_ISL_739908\| | ···················· | ···················· | ····················· | ···················· | ···················· | ···················· |
| Z | 2057 hCoV-19/Luxembourg/LNS1213113/2020\|EPI_ISL_739909\| | ···················· | ···················· | ····················· | ···················· | ···················· | ···················· |
| Z | 2058 hCoV-19/Luxembourg/LNS1239541/2020\|EPI_ISL_739910\| | ···················· | ···················· | ····················· | ···················· | ···················· | ···················· |
| Z | 2059 hCoV-19/Luxembourg/LNS1258846/2020\|EPI_ISL_739911\| | ···················· | ···················· | ····················· | ···················· | ···················· | ···················· |
| Z | 2060 hCoV-19/Luxembourg/LNS1271619/2020\|EPI_ISL_739912\| | ···················· | ···················· | ····················· | ···················· | ···················· | ···················· |
| Z | 2061 hCoV-19/Luxembourg/LNS1274513/2020\|EPI_ISL_739913\| | ···················· | ···················· | ····················· | ···················· | ···················· | ···················· |
| Z | 2062 hCoV-19/Luxembourg/LNS1290799/2020\|EPI_ISL_739914\| | ···················· | ···················· | ····················· | ···················· | ···················· | ···················· |
| Z | 2063 hCoV-19/Luxembourg/LNS1294821/2020\|EPI_ISL_739915\| | ···················· | ···················· | ····················· | ···················· | ···················· | ···················· |
| Z | 2064 hCoV-19/Luxembourg/LNS1301720/2020\|EPI_ISL_739916\| | ···················· | ···················· | ····················· | ···················· | ···················· | ···················· |
| Z | 2065 hCoV-19/Luxembourg/LNS1464616/2020\|EPI_ISL_739939\| | ···················· | ···················· | ····················· | ···················· | ···················· | ···················· |
| Z | 2066 hCoV-19/Luxembourg/LNS1527323/2020\|EPI_ISL_739948\| | ···················· | ···················· | ····················· | ···················· | ···················· | ···················· |
| Z | 2067 hCoV-19/Luxembourg/LNS1531431/2020\|EPI_ISL_739950\| | ···················· | ···················· | ····················· | ···················· | ···················· | ···················· |
| Z | 2068 hCoV-19/Luxembourg/LNS1539378/2020\|EPI_ISL_739952\| | ···················· | ···················· | ····················· | ···················· | ···················· | ···················· |
| Z | 2069 hCoV-19/Luxembourg/LNS1540979/2020\|EPI_ISL_739953\| | ···················· | ···················· | ····················· | ···················· | ···················· | ···················· |
| Z | 2070 hCoV-19/Luxembourg/LNS1553454/2020\|EPI_ISL_739954\| | ···················· | ····A··············· | ····················· | ···················· | ···················· | ···················· |
| Z | 2071 hCoV-19/Luxembourg/LNS1566975/2020\|EPI_ISL_739955\| | ···················· | ···················· | ····················· | ···················· | ······T············· | ···················· |
| Z | 2072 hCoV-19/Luxembourg/LNS1567917/2020\|EPI_ISL_739956\| | ···················· | ···················· | ····················· | ···················· | ···················· | ···················· |
| Z | 2073 hCoV-19/Luxembourg/LNS1571603/2020\|EPI_ISL_739957\| | ···················· | ···················· | ····················· | ···················· | ···················· | ···················· |
| Z | 2074 hCoV-19/Luxembourg/LNS1590999/2020\|EPI_ISL_739958\| | ···················· | ···················· | ····················· | ···················· | ···················· | ···················· |
| Z | 2075 hCoV-19/England/ALDP-B75821/2020\|EPI_ISL_740926\|20 | ···················· | ···················· | ····················· | ···················· | ···················· | ···················· |
| Z | 2076 hCoV-19/England/ALDP-B7584F/2020\|EPI_ISL_740927\|20 | ···················· | ···················· | ····················· | ···················· | ···················· | ···················· |
| Z | 2077 hCoV-19/England/ALDP-B759A6/2020\|EPI_ISL_740931\|20 | ···················· | ···················· | ····················· | ···················· | ···················· | ···················· |
| Z | 2078 hCoV-19/England/ALDP-B759B5/2020\|EPI_ISL_740932\|20 | ···················· | ···················· | ····················· | ···················· | ···················· | ···················· |
| Z | 2079 hCoV-19/England/ALDP-B759C4/2020\|EPI_ISL_740933\|20 | ···················· | ···················· | ····················· | ···················· | ···················· | ···················· |
| Z | 2080 hCoV-19/England/ALDP-B759D3/2020\|EPI_ISL_740934\|20 | ···················· | ···················· | ····················· | ···················· | ···················· | ···················· |
| Z | 2081 hCoV-19/England/ALDP-B75AFE/2020\|EPI_ISL_740935\|20 | ···················· | ···················· | ····················· | ···················· | ···················· | ···················· |
| Z | 2082 hCoV-19/England/ALDP-B75B0A/2020\|EPI_ISL_740936\|20 | ···················· | ···················· | ····················· | ···················· | ···················· | ···················· |
| Z | 2083 hCoV-19/England/ALDP-B75B37/2020\|EPI_ISL_740937\|20 | ···················· | ···················· | ····················· | ···················· | ······T············· | ···················· |
| Z | 2084 hCoV-19/England/ALDP-B75B73/2020\|EPI_ISL_740939\|20 | ···················· | ···················· | ····················· | ···················· | ···················· | ···················· |
| Z | 2085 hCoV-19/Wales/PHWC-49EBA8/2020\|EPI_ISL_743101\|2020 | ···················· | ···················· | ····················· | ···················· | ···················· | ···················· |
| Z | 2086 hCoV-19/Wales/PHWC-49EBC6/2020\|EPI_ISL_743103\|2020 | ···················· | ···················· | ····················· | ···················· | ···················· | ···················· |
| Z | 2087 hCoV-19/Wales/PHWC-49EC0F/2020\|EPI_ISL_743104\|2020 | ···················· | ···················· | ····················· | ···················· | ···················· | ···················· |
| Z | 2088 hCoV-19/Wales/PHWC-49EC2D/2020\|EPI_ISL_743105\|2020 | ···················· | ···················· | ····················· | ···················· | ···················· | ···················· |
| Z | 2089 hCoV-19/Wales/PHWC-49EC69/2020\|EPI_ISL_743107\|2020 | ···················· | ···················· | ····················· | ···················· | ···················· | ···················· |
| Z | 2091 hCoV-19/Wales/PHWC-49ECA5/2020\|EPI_ISL_743109\|2020 | ···················· | ···················· | ····················· | ···················· | ···················· | ···················· |
| Z | 2092 hCoV-19/Wales/PHWC-49ECC3/2020\|EPI_ISL_743110\|2020 | ···················· | ···················· | ····················· | ···················· | ···················· | ···················· |
| Z | 2094 hCoV-19/Wales/PHWC-49ED1B/2020\|EPI_ISL_743112\|2020 | ···················· | ···················· | ····················· | ···················· | ···················· | ···················· |
| Z | 2095 hCoV-19/Wales/PHWC-49ED2A/2020\|EPI_ISL_743113\|2020 | ···················· | ···················· | ····················· | ···················· | ···················· | ···················· |
| Z | 2096 hCoV-19/Wales/PHWC-49ED39/2020\|EPI_ISL_743114\|2020 | ···················· | ···················· | ····················· | ···················· | ···················· | ···················· |
| Z | 2097 hCoV-19/Wales/PHWC-49ED48/2020\|EPI_ISL_743115\|2020 | ···················· | ···················· | ····················· | ···················· | ···················· | ···················· |
| Z | 2100 hCoV-19/Wales/PHWC-49ED75/2020\|EPI_ISL_743118\|2020 | ···················· | ···················· | ····················· | ···················· | ···················· | ···················· |
| Z | 2102 hCoV-19/Wales/PHWC-49ED93/2020\|EPI_ISL_743120\|2020 | ···················· | ···················· | ····················· | ···················· | ···················· | ···················· |
| Z | 2103 hCoV-19/Wales/PHWC-49EDB1/2020\|EPI_ISL_743122\|2020 | ···················· | ···················· | ····················· | ···················· | ···················· | ···················· |
| Z | 2106 hCoV-19/Wales/PHWC-49EDEE/2020\|EPI_ISL_743125\|2020 | ···················· | ···················· | ····················· | ···················· | ···················· | ···················· |
| Z | 2108 hCoV-19/Wales/PHWC-49EE09/2020\|EPI_ISL_743127\|2020 | ···················· | ···················· | ····················· | ···················· | ···················· | ···················· |
| Z | 2110 hCoV-19/Wales/PHWC-49EE54/2020\|EPI_ISL_743132\|2020 | ···················· | ···················· | ····················· | ···················· | ···················· | ···················· |
| Z | 2113 hCoV-19/Wales/PHWC-49EE81/2020\|EPI_ISL_743135\|2020 | ···················· | ···················· | ····················· | ···················· | ···················· | ···················· |
| Z | 2115 hCoV-19/Wales/PHWC-49EEBE/2020\|EPI_ISL_743138\|2020 | ···················· | ···················· | ····················· | ···················· | ···················· | ···················· |
| Z | 2116 hCoV-19/Wales/PHWC-49EEEB/2020\|EPI_ISL_743140\|2020 | ···················· | ···················· | ····················· | ···················· | ···················· | ···················· |
| Z | 2117 hCoV-19/Wales/PHWC-49EEFA/2020\|EPI_ISL_743141\|2020 | ···················· | ···················· | ····················· | ···················· | ···················· | ···················· |
| Z | 2118 hCoV-19/Wales/PHWC-49EF06/2020\|EPI_ISL_743142\|2020 | ···················· | ···················· | ····················· | ···················· | ···················· | ···················· |
| Z | 2119 hCoV-19/Wales/PHWC-49EF15/2020\|EPI_ISL_743143\|2020 | ···················· | ···················· | ····················· | ···················· | ···················· | ···················· |
| Z | 2120 hCoV-19/Wales/PHWC-49EF24/2020\|EPI_ISL_743144\|2020 | ···················· | ···················· | ····················· | ···················· | ···················· | ···················· |
| Z | 2121 hCoV-19/Wales/PHWC-49EF33/2020\|EPI_ISL_743145\|2020 | ···················· | ···················· | ····················· | ···················· | ···················· | ···················· |
| Z | 2122 hCoV-19/Wales/PHWC-49EF42/2020\|EPI_ISL_743146\|2020 | ···················· | ···················· | ····················· | ···················· | ···················· | ···················· |
| Z | 2124 hCoV-19/Wales/PHWC-49EF7F/2020\|EPI_ISL_743149\|2020 | ···················· | ···················· | ····················· | ···················· | ···················· | ···················· |
| Z | 2125 hCoV-19/Wales/PHWC-49EF9D/2020\|EPI_ISL_743150\|2020 | ···················· | ···················· | ····················· | ···················· | ···················· | ···················· |
| Z | 2126 hCoV-19/Wales/PHWC-49EFBB/2020\|EPI_ISL_743151\|2020 | ···················· | ···················· | ····················· | ···················· | ···················· | ···················· |
| Z | 2131 hCoV-19/Wales/PHWC-49F020/2020\|EPI_ISL_743158\|2020 | ···················· | ···················· | ····················· | ···················· | ···················· | ···················· |
| Z | 2132 hCoV-19/Wales/PHWC-49F04E/2020\|EPI_ISL_743159\|2020 | ···················· | ···················· | ····················· | ···················· | ···················· | ···················· |
| Z | 2133 hCoV-19/Wales/PHWC-49F05D/2020\|EPI_ISL_743160\|2020 | ···················· | ···················· | ····················· | ···················· | ···················· | ···················· |
| Z | 2134 hCoV-19/Wales/PHWC-49F08A/2020\|EPI_ISL_743162\|2020 | ···················· | ···················· | ····················· | ···················· | ···················· | ···················· |
| Z | 2135 hCoV-19/Wales/PHWC-49F099/2020\|EPI_ISL_743163\|2020 | ···················· | ···················· | ····················· | ···················· | ···················· | ···················· |
| Z | 2136 hCoV-19/Wales/PHWC-49F0A8/2020\|EPI_ISL_743164\|2020 | ···················· | ···················· | ····················· | ···················· | ···················· | ···················· |
| Z | 2140 hCoV-19/Wales/PHWC-49F13C/2020\|EPI_ISL_743170\|2020 | ···················· | ···················· | ····················· | ···················· | ···················· | ···················· |
| Z | 2142 hCoV-19/Wales/PHWC-49F15A/2020\|EPI_ISL_743172\|2020 | ···················· | ···················· | ····················· | ···················· | ···················· | ···················· |
| Z | 2144 hCoV-19/Wales/PHWC-49F196/2020\|EPI_ISL_743174\|2020 | ···················· | ···················· | ····················· | ···················· | ···················· | ···················· |
| Z | 2145 hCoV-19/Wales/PHWC-49F1A5/2020\|EPI_ISL_743175\|2020 | ···················· | ···················· | ····················· | ···················· | ···················· | ···················· |
| Z | 2146 hCoV-19/Wales/PHWC-49F1B4/2020\|EPI_ISL_743176\|2020 | ···················· | ···················· | ····················· | ···················· | ···················· | ···················· |
| Z | 2147 hCoV-19/Wales/PHWC-49F1C3/2020\|EPI_ISL_743177\|2020 | ···················· | ···················· | ····················· | ···················· | ···················· | ···················· |
| Z | 2149 hCoV-19/Wales/PHWC-49F1E1/2020\|EPI_ISL_743179\|2020 | ···················· | ···················· | ····················· | ···················· | ···················· | ···················· |
| Z | 2150 hCoV-19/Luxembourg/LNS6346257/2020\|EPI_ISL_744400\| | ···················· | ···················· | ····················· | ···················· | ···················· | ···················· |
| Z | 2151 hCoV-19/Luxembourg/LNS6356413/2020\|EPI_ISL_744401\| | ···················· | ···················· | ····················· | ···················· | ···················· | ···················· |
| Z | 2152 hCoV-19/Luxembourg/LNS6358751/2020\|EPI_ISL_744402\| | ···················· | ···················· | ····················· | ···················· | ···················· | ···················· |

| | | | | | | | | |
|---|---|---|---|---|---|---|---|---|
| Z | 2153 | hCoV-19/Luxembourg/LNS6379157/2020\|EPI_ISL_744404\| | ···················· | ···················· | ····················· | ···················· | ···················· | ···················· |
| Z | 2154 | hCoV-19/Luxembourg/LNS6379862/2020\|EPI_ISL_744405\| | ···················· | ···················· | ····················· | ···················· | ······T············· | ···················· |
| Z | 2155 | hCoV-19/Luxembourg/LNS6380996/2020\|EPI_ISL_744406\| | ···················· | ···················· | ····················· | ···················· | ···················· | ···················· |
| Z | 2156 | hCoV-19/Luxembourg/LNS6409176/2020\|EPI_ISL_744407\| | ···················· | ···················· | ····················· | ···················· | ···················· | ···················· |
| Z | 2157 | hCoV-19/Luxembourg/LNS6417532/2020\|EPI_ISL_744408\| | ···················· | ···················· | ····················· | ···················· | ···················· | ···················· |
| Z | 2158 | hCoV-19/Luxembourg/LNS6422627/2020\|EPI_ISL_744409\| | ···················· | ···················· | ····················· | ···················· | ···················· | ···················· |
| Z | 2159 | hCoV-19/Luxembourg/LNS6429095/2020\|EPI_ISL_744411\| | ···················· | ···················· | ····················· | ···················· | ···················· | ···················· |
| Z | 2160 | hCoV-19/Luxembourg/LNS6429244/2020\|EPI_ISL_744412\| | ···················· | ···················· | ····················· | ···················· | ···················· | ···················· |
| Z | 2161 | hCoV-19/Luxembourg/LNS6434733/2020\|EPI_ISL_744413\| | ···················· | ···················· | ····················· | ···················· | ······T············· | ···················· |
| Z | 2162 | hCoV-19/Luxembourg/LNS6437250/2020\|EPI_ISL_744414\| | ···················· | ···················· | ····················· | ···················· | ···················· | ···················· |
| Z | 2163 | hCoV-19/Luxembourg/LNS6445912/2020\|EPI_ISL_744415\| | ···················· | ···················· | ····················· | ···················· | ···················· | ···················· |
| Z | 2164 | hCoV-19/Luxembourg/LNS6527887/2020\|EPI_ISL_744427\| | ···················· | G··················· | ····················· | ···················· | ···················· | ···················· |
| Z | 2165 | hCoV-19/Luxembourg/LNS6539305/2020\|EPI_ISL_744428\| | ···················· | ···················· | ····················· | ···················· | ···················· | ···················· |
| Z | 2166 | hCoV-19/Luxembourg/LNS6546741/2020\|EPI_ISL_744429\| | ···········T········ | ···················· | ····················· | ···················· | ···················· | ···················· |
| Z | 2167 | hCoV-19/Luxembourg/LNS6569914/2020\|EPI_ISL_744430\| | ···················· | ···················· | ····················· | ···················· | ···················· | ···················· |
| Z | 2168 | hCoV-19/Luxembourg/LNS6570261/2020\|EPI_ISL_744431\| | ···················· | ···················· | ····················· | ···················· | ···················· | ···················· |
| Z | 2169 | hCoV-19/Luxembourg/LNS6572463/2020\|EPI_ISL_744432\| | ···················· | ···················· | ····················· | ···················· | ···················· | ···················· |
| Z | 2170 | hCoV-19/Luxembourg/LNS6578763/2020\|EPI_ISL_744433\| | ···················· | ···················· | ····················· | ···················· | ···················· | ···················· |
| Z | 2171 | hCoV-19/Luxembourg/LNS6579692/2020\|EPI_ISL_744434\| | ···················· | ···················· | ····················· | ···················· | ···················· | ···················· |
| Z | 2172 | hCoV-19/Luxembourg/LNS6582736/2020\|EPI_ISL_744435\| | ···················· | ···················· | ····················· | ···················· | ···················· | ···················· |
| Z | 2173 | hCoV-19/Luxembourg/LNS6593260/2020\|EPI_ISL_744436\| | ···················· | ···················· | ····················· | ···················· | ···················· | ···················· |
| Z | 2174 | hCoV-19/Luxembourg/LNS6596752/2020\|EPI_ISL_744437\| | ···················· | ···················· | ····················· | ···················· | ···················· | ···················· |
| Z | 2175 | hCoV-19/Luxembourg/LNS6599503/2020\|EPI_ISL_744438\| | ···················· | ···················· | ····················· | ···················· | ···················· | ···················· |
| Z | 2176 | hCoV-19/Luxembourg/LNS6600836/2020\|EPI_ISL_744439\| | ···················· | ··········A········· | ····················· | ···················· | ···················· | ···················· |
| Z | 2177 | hCoV-19/Luxembourg/LNS6608981/2020\|EPI_ISL_744440\| | ···················· | ···················· | ····················· | ···················· | ···················· | ···················· |
| Z | 2178 | hCoV-19/Luxembourg/LNS6618380/2020\|EPI_ISL_744441\| | ···················· | ···················· | ····················· | ···················· | ···················· | ···················· |
| Z | 2179 | hCoV-19/Luxembourg/LNS6619182/2020\|EPI_ISL_744442\| | ···················· | ···················· | ····················· | ···················· | ···················· | ···················· |
| Z | 2180 | hCoV-19/Luxembourg/LNS6636441/2020\|EPI_ISL_744443\| | ···················· | ···················· | ····················· | ···················· | ···················· | ···················· |
| Z | 2181 | hCoV-19/Luxembourg/LNS6650712/2020\|EPI_ISL_744444\| | ···················· | ···················· | ····················· | ···················· | ···················· | ···················· |
| Z | 2182 | hCoV-19/Luxembourg/LNS6652363/2020\|EPI_ISL_744445\| | ···················· | ···················· | ····················· | ···················· | ······T············· | ···················· |
| Z | 2183 | hCoV-19/Luxembourg/LNS6663810/2020\|EPI_ISL_744446\| | ···················· | ···················· | ····················· | ···················· | ···················· | ···················· |
| Z | 2184 | hCoV-19/Luxembourg/LNS6665496/2020\|EPI_ISL_744447\| | ···················· | ···················· | ····················· | ···················· | ···················· | ···················· |
| Z | 2185 | hCoV-19/Luxembourg/LNS6665821/2020\|EPI_ISL_744448\| | ···················· | ···················· | ····················· | ···················· | ···················· | ···················· |
| Z | 2186 | hCoV-19/Luxembourg/LNS6686752/2020\|EPI_ISL_744449\| | ···················· | ···················· | ····················· | ···················· | ······T············· | ···················· |
| Z | 2187 | hCoV-19/Luxembourg/LNS6688631/2020\|EPI_ISL_744450\| | ···················· | ···················· | ····················· | ···················· | ···················· | ···················· |
| Z | 2188 | hCoV-19/Luxembourg/LNS6691015/2020\|EPI_ISL_744451\| | ···················· | ···················· | ····················· | ···················· | ···················· | ···················· |
| Z | 2189 | hCoV-19/Luxembourg/LNS6694971/2020\|EPI_ISL_744452\| | ···················· | ···················· | ····················· | ···················· | ···················· | ···················· |
| Z | 2190 | hCoV-19/Luxembourg/LNS6705842/2020\|EPI_ISL_744453\| | ···················· | ·A·················· | ····················· | ···················· | ···················· | ···················· |
| Z | 2191 | hCoV-19/Luxembourg/LNS6708829/2020\|EPI_ISL_744454\| | ···················· | ···················· | ····················· | ···················· | ···················· | ···················· |
| Z | 2192 | hCoV-19/Luxembourg/LNS6716560/2020\|EPI_ISL_744455\| | ···················· | ···················· | ····················· | ···················· | ···················· | ···················· |
| Z | 2193 | hCoV-19/Luxembourg/LNS6721228/2020\|EPI_ISL_744456\| | ···················· | ···················· | ····················· | ···················· | ···················· | ···················· |
| Z | 2194 | hCoV-19/Luxembourg/LNS6722271/2020\|EPI_ISL_744457\| | ···················· | ···················· | ····················· | ···················· | ···················· | ···················· |
| Z | 2195 | hCoV-19/Luxembourg/LNS6723123/2020\|EPI_ISL_744458\| | ···················· | ···················· | ····················· | ···················· | ···················· | ···················· |
| Z | 2196 | hCoV-19/Luxembourg/LNS6734577/2020\|EPI_ISL_744459\| | ···················· | ···················· | ····················· | ···················· | ···················· | ···················· |
| Z | 2197 | hCoV-19/Luxembourg/LNS6741137/2020\|EPI_ISL_744460\| | ···················· | ···················· | ····················· | ···················· | ···················· | ···················· |
| Z | 2198 | hCoV-19/Luxembourg/LNS6743711/2020\|EPI_ISL_744461\| | ···················· | ···················· | ····················· | ···················· | ···················· | ···················· |
| Z | 2199 | hCoV-19/Luxembourg/LNS6744641/2020\|EPI_ISL_744462\| | ···················· | ···················· | ····················· | ···················· | ···················· | ···················· |
| Z | 2200 | hCoV-19/Luxembourg/LNS6748939/2020\|EPI_ISL_744463\| | ···················· | ···················· | ····················· | ···················· | ···················· | ···················· |
| Z | 2201 | hCoV-19/Luxembourg/LNS6755246/2020\|EPI_ISL_744465\| | ············T······· | ···················· | ····················· | ···················· | ···················· | ···················· |
| Z | 2202 | hCoV-19/Luxembourg/LNS6762167/2020\|EPI_ISL_744466\| | ···················· | ···················· | ····················· | ···················· | ···················· | ···················· |
| Z | 2203 | hCoV-19/Luxembourg/LNS6766642/2020\|EPI_ISL_744467\| | ···················· | ···················· | ····················· | ···················· | ···················· | ···················· |
| Z | 2204 | hCoV-19/Luxembourg/LNS6772428/2020\|EPI_ISL_744468\| | ···················· | ···················· | ····················· | ···················· | ···················· | ···················· |
| Z | 2205 | hCoV-19/Luxembourg/LNS6773403/2020\|EPI_ISL_744469\| | ···················· | ···················· | ····················· | ···················· | ···················· | ···················· |
| Z | 2206 | hCoV-19/Luxembourg/LNS6777622/2020\|EPI_ISL_744470\| | ···················· | ···················· | ····················· | ···················· | ···················· | ···················· |
| Z | 2207 | hCoV-19/Luxembourg/LNS6777691/2020\|EPI_ISL_744471\| | ···················· | ···················· | ····················· | ···················· | ······T············· | ···················· |
| Z | 2208 | hCoV-19/Luxembourg/LNS6807787/2020\|EPI_ISL_744472\| | ···················· | ···················· | ····················· | ···················· | ···················· | ···················· |
| Z | 2209 | hCoV-19/Luxembourg/LNS6812319/2020\|EPI_ISL_744473\| | ···················· | ···················· | ····················· | ···················· | ···················· | ···················· |
| Z | 2210 | hCoV-19/Luxembourg/LNS6839721/2020\|EPI_ISL_744474\| | ···················· | ···················· | ····················· | ···················· | ···················· | ···················· |
| Z | 2211 | hCoV-19/Luxembourg/LNS6858142/2020\|EPI_ISL_744475\| | ···················· | ···················· | ····················· | ···················· | ···················· | ···················· |
| Z | 2212 | hCoV-19/Luxembourg/LNS6878538/2020\|EPI_ISL_744476\| | ···················· | ···················· | ····················· | ···················· | ···················· | ···················· |
| Z | 2213 | hCoV-19/Luxembourg/LNS6883359/2020\|EPI_ISL_744478\| | ···················· | ···················· | ····················· | ···················· | ···················· | ···················· |
| Z | 2214 | hCoV-19/Luxembourg/LNS6888018/2020\|EPI_ISL_744479\| | ···················· | ···················· | ····················· | ···················· | ···················· | ···················· |
| Z | 2215 | hCoV-19/Luxembourg/LNS6889202/2020\|EPI_ISL_744480\| | ···················· | ···················· | ····················· | ···················· | ···················· | ···················· |
| Z | 2216 | hCoV-19/Luxembourg/LNS6894146/2020\|EPI_ISL_744481\| | ···················· | ···················· | ····················· | ···················· | ···················· | ···················· |
| Z | 2217 | hCoV-19/Luxembourg/LNS6898993/2020\|EPI_ISL_744482\| | ···················· | ···················· | ····················· | ···················· | ···················· | ···················· |
| Z | 2218 | hCoV-19/Luxembourg/LNS6906942/2020\|EPI_ISL_744483\| | ···················· | ···················· | ····················· | ···················· | ···················· | ···················· |
| Z | 2219 | hCoV-19/Luxembourg/LNS6959492/2020\|EPI_ISL_744486\| | ···················· | ···················· | ····················· | ···················· | ···················· | ···················· |
| Z | 2220 | hCoV-19/Luxembourg/LNS6961161/2020\|EPI_ISL_744487\| | ···················· | ···················· | ····················· | ···················· | ···················· | ···················· |
| Z | 2221 | hCoV-19/Luxembourg/LNS6975153/2020\|EPI_ISL_744489\| | ···················· | ···················· | ····················· | ···················· | ···················· | ···················· |
| Z | 2222 | hCoV-19/Luxembourg/LNS6976789/2020\|EPI_ISL_744490\| | ···················· | ···················· | ····················· | ···················· | ···················· | ···················· |
| Z | 2223 | hCoV-19/Luxembourg/LNS6978482/2020\|EPI_ISL_744491\| | ···················· | ···················· | ····················· | ···················· | ···················· | ···················· |
| Z | 2224 | hCoV-19/Luxembourg/LNS6982840/2020\|EPI_ISL_744492\| | ···················· | ···················· | ····················· | ···················· | ···················· | ···················· |
| Z | 2225 | hCoV-19/Luxembourg/LNS6986022/2020\|EPI_ISL_744493\| | ···················· | ···················· | ····················· | ···················· | ···················· | ···················· |
| Z | 2226 | hCoV-19/Luxembourg/LNS7022254/2020\|EPI_ISL_744494\| | ···················· | ···················· | ····················· | ···················· | ···················· | ···················· |
| Z | 2227 | hCoV-19/Luxembourg/LNS7025344/2020\|EPI_ISL_744495\| | ···················· | ···················· | ····················· | ···················· | ···················· | ···················· |
| Z | 2228 | hCoV-19/Luxembourg/LNS7026792/2020\|EPI_ISL_744496\| | ···················· | ···················· | ····················· | ···················· | ···················· | ···················· |
| Z | 2229 | hCoV-19/Luxembourg/LNS7028695/2020\|EPI_ISL_744497\| | ···················· | ···················· | ····················· | ···················· | ···················· | ···················· |
| Z | 2230 | hCoV-19/Luxembourg/LNS7028861/2020\|EPI_ISL_744498\| | ···················· | ···················· | ····················· | ···················· | ···················· | ···················· |
| Z | 2231 | hCoV-19/Luxembourg/LNS7030774/2020\|EPI_ISL_744499\| | ···················· | ···················· | ····················· | ···················· | ···················· | ···················· |
| Z | 2232 | hCoV-19/USA/UT-UPHL-2012736905/2020\|EPI_ISL_745714 | ···················· | ···················· | ····················· | ···················· | ···················· | ···················· |

| | | | | | | | | |
|---|---|---|---|---|---|---|---|---|
| Z | 2233 | hCoV-19/USA/UT-UPHL-2012529719/2020\|EPI_ISL_745715 | ···················· | ·A·················· | ····················· | ···················· | ···················· | ···················· |
| Z | 2234 | hCoV-19/USA/UT-UPHL-2012070308/2020\|EPI_ISL_745716 | ···················· | ·A·················· | ····················· | ···················· | ···················· | ···················· |
| Z | 2235 | hCoV-19/USA/UT-UPHL-2012453480/2020\|EPI_ISL_745717 | ···················· | ···················· | ····················· | ···················· | ···················· | ···················· |
| Z | 2236 | hCoV-19/USA/UT-UPHL-2012266098/2020\|EPI_ISL_745718 | ···················· | ·A·················· | ····················· | ···················· | ···················· | ···················· |
| Z | 2237 | hCoV-19/USA/UT-UPHL-2012029219/2020\|EPI_ISL_745719 | ···················· | ···················· | ····················· | ···················· | ·········A·········· | ···················· |
| Z | 2238 | hCoV-19/USA/UT-UPHL-2012289921/2020\|EPI_ISL_745720 | ···················· | ···················· | ····················· | ···················· | ···················· | ···················· |
| Z | 2239 | hCoV-19/USA/UT-UPHL-2012379451/2020\|EPI_ISL_745721 | ···················· | ···················· | ····················· | ···················· | ···················· | ···················· |
| Z | 2240 | hCoV-19/USA/UT-UPHL-2012652261/2020\|EPI_ISL_745722 | ···················· | ···················· | ····················· | ···················· | ···················· | ···················· |
| Z | 2241 | hCoV-19/USA/UT-UPHL-2012620376/2020\|EPI_ISL_745723 | ···················· | ·A·················· | ····················· | ···················· | ···················· | ···················· |
| Z | 2242 | hCoV-19/USA/UT-UPHL-2012183575/2020\|EPI_ISL_745724 | ···················· | ···················· | ····················· | ···················· | ···················· | ···················· |
| Z | 2243 | hCoV-19/USA/UT-UPHL-2012061461/2020\|EPI_ISL_745725 | ···················· | ···················· | ····················· | ···················· | ···················· | ···················· |
| Z | 2244 | hCoV-19/USA/UT-UPHL-2012830401/2020\|EPI_ISL_745726 | ···················· | ···················· | ····················· | ···················· | ···················· | ···················· |
| Z | 2245 | hCoV-19/USA/UT-UPHL-2012322333/2020\|EPI_ISL_745727 | ···················· | ···················· | ····················· | ···················· | ···················· | ···················· |
| Z | 2246 | hCoV-19/USA/UT-UPHL-2012062404/2020\|EPI_ISL_745728 | ···················· | ·A·················· | ····················· | ···················· | ···················· | ···················· |
| Z | 2247 | hCoV-19/USA/UT-UPHL-2012824129/2020\|EPI_ISL_745729 | ···················· | ·A·················· | ····················· | ···················· | ···················· | ···················· |
| Z | 2248 | hCoV-19/USA/UT-UPHL-2012645552/2020\|EPI_ISL_745730 | ···················· | ·A·················· | ····················· | ···················· | ···················· | ···················· |
| Z | 2249 | hCoV-19/USA/UT-UPHL-2012590673/2020\|EPI_ISL_745731 | ···················· | ···················· | ····················· | ···················· | ···················· | ···················· |
| Z | 2250 | hCoV-19/USA/UT-UPHL-2012067509/2020\|EPI_ISL_745732 | ···················· | ···················· | ····················· | ···················· | ···················· | ···················· |
| Z | 2251 | hCoV-19/USA/UT-UPHL-2012329672/2020\|EPI_ISL_745733 | ···················· | ·A·················· | ····················· | ···················· | ···················· | ···················· |
| Z | 2252 | hCoV-19/USA/UT-UPHL-2012611872/2020\|EPI_ISL_745734 | ···················· | ···················· | ····················· | ···················· | ···················· | ···················· |
| Z | 2253 | hCoV-19/USA/UT-UPHL-2012625809/2020\|EPI_ISL_745735 | ···················· | ···················· | ····················· | ···················· | ···················· | ···················· |
| Z | 2254 | hCoV-19/USA/UT-UPHL-2012150388/2020\|EPI_ISL_745736 | ···················· | ···················· | ····················· | ···················· | ···················· | ···················· |
| Z | 2255 | hCoV-19/USA/UT-UPHL-2012200340/2020\|EPI_ISL_745737 | ···················· | ···················· | ····················· | ···················· | ···················· | ···················· |
| Z | 2256 | hCoV-19/USA/UT-UPHL-2012772719/2020\|EPI_ISL_745738 | ···················· | ···················· | ····················· | ···················· | ···················· | ···················· |
| Z | 2257 | hCoV-19/USA/UT-UPHL-2012459187/2020\|EPI_ISL_745739 | ···················· | ···················· | ····················· | ···················· | ···················· | ···················· |
| Z | 2258 | hCoV-19/USA/UT-UPHL-2012861567/2020\|EPI_ISL_745740 | ···················· | ···················· | ····················· | ···················· | ···················· | ···················· |
| Z | 2259 | hCoV-19/USA/UT-UPHL-2012937963/2020\|EPI_ISL_745743 | ···················· | ···················· | ····················· | ···················· | ···················· | ···················· |
| Z | 2260 | hCoV-19/USA/UT-UPHL-2012874225/2020\|EPI_ISL_745744 | ···················· | ···················· | ····················· | ···················· | ···················· | ···················· |
| Z | 2261 | hCoV-19/USA/UT-UPHL-2012868180/2020\|EPI_ISL_745745 | ···················· | ···················· | ····················· | ···················· | ···················· | ···················· |
| Z | 2262 | hCoV-19/USA/UT-UPHL-2012803695/2020\|EPI_ISL_745746 | ···················· | ···················· | ····················· | ···················· | ···················· | ···················· |
| Z | 2263 | hCoV-19/USA/UT-UPHL-2012527847/2020\|EPI_ISL_745758 | ···················· | ···················· | ····················· | ···················· | ···················· | ···················· |
| Z | 2264 | hCoV-19/USA/UT-UPHL-2012252797/2020\|EPI_ISL_745759 | ···················· | ···················· | ····················· | ···················· | ···················· | ···················· |
| Z | 2265 | hCoV-19/USA/UT-UPHL-2012390126/2020\|EPI_ISL_745760 | ···················· | ···················· | ····················· | ···················· | ···················· | ···················· |
| Z | 2266 | hCoV-19/USA/UT-UPHL-2012876311/2020\|EPI_ISL_745761 | ···················· | ···················· | ····················· | ···················· | ···················· | ···················· |
| Z | 2267 | hCoV-19/USA/UT-UPHL-2012523265/2020\|EPI_ISL_745762 | ···················· | ···················· | ····················· | ···················· | ···················· | ···················· |
| Z | 2268 | hCoV-19/USA/UT-UPHL-2012979596/2020\|EPI_ISL_745763 | ···················· | ·A·················· | ····················· | ···················· | ···················· | ···················· |
| Z | 2269 | hCoV-19/USA/UT-UPHL-2012426357/2020\|EPI_ISL_745764 | ···················· | ···················· | ····················· | ···················· | ···················· | ···················· |
| Z | 2270 | hCoV-19/USA/UT-UPHL-2012250398/2020\|EPI_ISL_745765 | ···················· | ···················· | ····················· | ···················· | ···················· | ···················· |
| Z | 2271 | hCoV-19/USA/UT-UPHL-2012444865/2020\|EPI_ISL_745766 | ···················· | ···················· | ····················· | ···················· | ···················· | ···················· |
| Z | 2272 | hCoV-19/USA/UT-UPHL-2012575394/2020\|EPI_ISL_745767 | ···················· | ···················· | ····················· | ···················· | ···················· | ···················· |
| Z | 2273 | hCoV-19/USA/UT-UPHL-2012828260/2020\|EPI_ISL_745768 | ···················· | ·A·················· | ····················· | ···················· | ···················· | ···················· |
| Z | 2274 | hCoV-19/USA/UT-UPHL-2012282366/2020\|EPI_ISL_745769 | ···················· | ···················· | ····················· | ···················· | ···················· | ···················· |
| Z | 2275 | hCoV-19/USA/UT-UPHL-2012270978/2020\|EPI_ISL_745770 | ···················· | ···················· | ····················· | ···················· | ···················· | ···················· |
| Z | 2276 | hCoV-19/USA/UT-UPHL-2012403274/2020\|EPI_ISL_745771 | ···················· | ·A·················· | ····················· | ···················· | ···················· | ···················· |
| Z | 2277 | hCoV-19/USA/UT-UPHL-2012695241/2020\|EPI_ISL_745772 | ···················· | ···················· | ····················· | ···················· | ···················· | ···················· |
| Z | 2278 | hCoV-19/USA/UT-UPHL-2012747464/2020\|EPI_ISL_745773 | ···················· | ···················· | ····················· | ···················· | ···················· | ···················· |
| Z | 2279 | hCoV-19/USA/UT-UPHL-2012491321/2020\|EPI_ISL_745774 | ···················· | ·A·················· | ····················· | ···················· | ···················· | ···················· |
| Z | 2280 | hCoV-19/USA/UT-UPHL-2012885604/2020\|EPI_ISL_745775 | ···················· | ·A·················· | ····················· | ···················· | ···················· | ···················· |
| Z | 2281 | hCoV-19/USA/UT-UPHL-2012973983/2020\|EPI_ISL_745776 | ···················· | ···················· | ····················· | ···················· | ···················· | ···················· |
| Z | 2282 | hCoV-19/USA/UT-UPHL-2012991735/2020\|EPI_ISL_745777 | ···················· | ···················· | ····················· | ···················· | ···················· | ···················· |
| Z | 2283 | hCoV-19/USA/UT-UPHL-2012614422/2020\|EPI_ISL_745778 | ···················· | ···················· | ····················· | ···················· | ···················· | ···················· |
| Z | 2284 | hCoV-19/USA/UT-UPHL-2012298356/2020\|EPI_ISL_745779 | ···················· | ·A·················· | ····················· | ···················· | ···················· | ···················· |
| Z | 2285 | hCoV-19/USA/UT-UPHL-2012777664/2020\|EPI_ISL_745780 | ···················· | ···················· | ····················· | ···················· | ···················· | ···················· |
| Z | 2286 | hCoV-19/USA/UT-UPHL-2012018896/2020\|EPI_ISL_745781 | ···················· | ·A·················· | ····················· | ···················· | ···················· | ···················· |
| Z | 2287 | hCoV-19/USA/UT-UPHL-2012474856/2020\|EPI_ISL_745782 | ···················· | ·A·················· | ····················· | ···················· | ···················· | ···················· |
| Z | 2288 | hCoV-19/USA/UT-UPHL-2012562796/2020\|EPI_ISL_745783 | ···················· | ·A·················· | ····················· | ···················· | ···················· | ···················· |
| Z | 2289 | hCoV-19/USA/UT-UPHL-2012118130/2020\|EPI_ISL_745784 | ···················· | ···················· | ····················· | ···················· | ···················· | ···················· |
| Z | 2290 | hCoV-19/USA/UT-UPHL-2012899488/2020\|EPI_ISL_745785 | ···················· | ···················· | ····················· | ···················· | ···················· | ···················· |
| Z | 2291 | hCoV-19/USA/UT-UPHL-2012189893/2020\|EPI_ISL_745786 | ···················· | ·A·················· | ····················· | ···················· | ···················· | ···················· |
| Z | 2292 | hCoV-19/USA/UT-UPHL-2012105256/2020\|EPI_ISL_745787 | ···················· | ···················· | ····················· | ···················· | ···················· | ···················· |
| Z | 2293 | hCoV-19/USA/UT-UPHL-2012623419/2020\|EPI_ISL_745788 | ···················· | ···················· | ····················· | ···················· | ···················· | ···················· |
| Z | 2294 | hCoV-19/USA/UT-UPHL-2012030607/2020\|EPI_ISL_745789 | ···················· | ···················· | ····················· | ···················· | ···················· | ···················· |
| Z | 2295 | hCoV-19/USA/UT-UPHL-2012811862/2020\|EPI_ISL_745790 | ···················· | ···················· | ····················· | ···················· | ···················· | ···················· |
| Z | 2296 | hCoV-19/USA/UT-UPHL-2012022396/2020\|EPI_ISL_745791 | ···················· | ···················· | ····················· | ···················· | ···················· | ···················· |
| Z | 2297 | hCoV-19/USA/UT-UPHL-2012534028/2020\|EPI_ISL_745792 | ···················· | ···················· | ····················· | ···················· | ···················· | ···················· |
| Z | 2298 | hCoV-19/USA/UT-UPHL-2012102790/2020\|EPI_ISL_745793 | ···················· | ···················· | ····················· | ···················· | ···················· | ···················· |
| Z | 2299 | hCoV-19/USA/UT-UPHL-2012077077/2020\|EPI_ISL_745794 | ···················· | ···················· | ····················· | ···················· | ···················· | ···················· |
| Z | 2300 | hCoV-19/USA/UT-UPHL-2012085032/2020\|EPI_ISL_745795 | ···················· | ·A·················· | ····················· | ···················· | ···················· | ···················· |
| Z | 2301 | hCoV-19/USA/UT-UPHL-2012099424/2020\|EPI_ISL_745796 | ···················· | ···················· | ····················· | ···················· | ···················· | ···················· |
| Z | 2302 | hCoV-19/USA/UT-UPHL-2012105352/2020\|EPI_ISL_745797 | ···················· | ·A·················· | ····················· | ···················· | ···················· | ···················· |
| Z | 2303 | hCoV-19/USA/UT-UPHL-2012023081/2020\|EPI_ISL_745798 | ···················· | ···················· | ····················· | ···················· | ···················· | ···················· |
| Z | 2304 | hCoV-19/USA/UT-UPHL-2012406252/2020\|EPI_ISL_745799 | ···················· | ···················· | ····················· | ···················· | ···················· | ···················· |
| Z | 2306 | hCoV-19/Luxembourg/LNS1306074/2020\|EPI_ISL_739917\| | ···················· | ···················· | ····················· | ···················· | ···················· | ···················· |
| Z | 2307 | hCoV-19/Luxembourg/LNS1308805/2020\|EPI_ISL_739918\| | ···················· | ···················· | ····················· | ···················· | ···················· | ···················· |
| Z | 2308 | hCoV-19/Luxembourg/LNS1316926/2020\|EPI_ISL_739919\| | ···················· | ···················· | ····················· | ···················· | ···················· | ···················· |
| Z | 2309 | hCoV-19/Luxembourg/LNS1340860/2020\|EPI_ISL_739921\| | ···················· | ···················· | ····················· | ···················· | ······T············· | ···················· |
| Z | 2310 | hCoV-19/Luxembourg/LNS1355488/2020\|EPI_ISL_739922\| | ···················· | ···················· | ····················· | ···················· | ···················· | ···················· |
| Z | 2311 | hCoV-19/Luxembourg/LNS1360776/2020\|EPI_ISL_739923\| | ···················· | ···················· | ····················· | ···················· | ···················· | ···················· |
| Z | 2312 | hCoV-19/Luxembourg/LNS1373805/2020\|EPI_ISL_739924\| | ···················· | ···················· | ····················· | ···················· | ···················· | ···················· |
| Z | 2313 | hCoV-19/Luxembourg/LNS1378944/2020\|EPI_ISL_739925\| | ···················· | ···················· | ····················· | ···················· | ······T············· | ···················· |

| | | | | | | | | |
|---|---|---|---|---|---|---|---|---|
| Z | 2314 | hCoV-19/Luxembourg/LNS1389060/2020\|EPI_ISL_739926\| | ···················· | ·A·················· | ····················· | ···················· | ···················· | ···················· |
| Z | 2315 | hCoV-19/Luxembourg/LNS1389642/2020\|EPI_ISL_739927\| | ···················· | ···················· | ····················· | ···················· | ···················· | ···················· |
| Z | 2316 | hCoV-19/Luxembourg/LNS1397730/2020\|EPI_ISL_739928\| | ···················· | ···················· | ····················· | ···················· | ···················· | ···················· |
| Z | 2317 | hCoV-19/Luxembourg/LNS1406195/2020\|EPI_ISL_739929\| | ···················· | ···················· | ····················· | ···················· | ···················· | ···················· |
| Z | 2318 | hCoV-19/Luxembourg/LNS1410268/2020\|EPI_ISL_739930\| | ···················· | ···················· | ····················· | ···················· | ···················· | ···················· |
| Z | 2319 | hCoV-19/Luxembourg/LNS1411791/2020\|EPI_ISL_739931\| | ···················· | ···················· | ····················· | ···················· | ···················· | ···················· |
| Z | 2320 | hCoV-19/Luxembourg/LNS1415676/2020\|EPI_ISL_739932\| | ···················· | ···················· | ····················· | ···················· | ···················· | ···················· |
| Z | 2321 | hCoV-19/Luxembourg/LNS1419064/2020\|EPI_ISL_739933\| | ···················· | ···················· | ····················· | ···················· | ···················· | ···················· |
| Z | 2322 | hCoV-19/Luxembourg/LNS1430659/2020\|EPI_ISL_739934\| | ···················· | ···················· | ····················· | ···················· | ···················· | ···················· |
| Z | 2323 | hCoV-19/Luxembourg/LNS1431754/2020\|EPI_ISL_739935\| | ···················· | ···················· | ····················· | ···················· | ···················· | ···················· |
| Z | 2324 | hCoV-19/Luxembourg/LNS1432866/2020\|EPI_ISL_739936\| | ···················· | ···················· | ···········T········· | ···················· | ···················· | ···················· |
| Z | 2325 | hCoV-19/Luxembourg/LNS1443623/2020\|EPI_ISL_739937\| | ···················· | ···················· | ····················· | ···················· | ···················· | ···················· |
| Z | 2326 | hCoV-19/Luxembourg/LNS1459568/2020\|EPI_ISL_739938\| | ···················· | ···················· | ····················· | ···················· | ···················· | ···················· |
| Z | 2327 | hCoV-19/Luxembourg/LNS1476390/2020\|EPI_ISL_739940\| | ···················· | ···················· | ····················· | ···················· | ···················· | ···················· |
| Z | 2328 | hCoV-19/Luxembourg/LNS1480431/2020\|EPI_ISL_739941\| | ···················· | ···················· | ····················· | ···················· | ···················· | ···················· |
| Z | 2329 | hCoV-19/Luxembourg/LNS1481285/2020\|EPI_ISL_739942\| | ···················· | ···················· | ····················· | ···················· | ···················· | ···················· |
| Z | 2330 | hCoV-19/Luxembourg/LNS1483758/2020\|EPI_ISL_739943\| | ···················· | ···················· | ····················· | ···················· | ···················· | ···················· |
| Z | 2331 | hCoV-19/Luxembourg/LNS1488367/2020\|EPI_ISL_739944\| | ···················· | ···················· | ····················· | ···················· | ···················· | ···················· |
| Z | 2332 | hCoV-19/Luxembourg/LNS1491016/2020\|EPI_ISL_739945\| | ···················· | ···················· | ····················· | ···················· | ···················· | ···················· |
| Z | 2333 | hCoV-19/Luxembourg/LNS1526149/2020\|EPI_ISL_739946\| | ···················· | ···················· | ····················· | ···················· | ···················· | ···················· |
| Z | 2334 | hCoV-19/Luxembourg/LNS1530163/2020\|EPI_ISL_739949\| | ···················· | ···················· | ····················· | ···················· | ···················· | ···················· |
| Z | 2335 | hCoV-19/Luxembourg/LNS1611085/2020\|EPI_ISL_739960\| | ···················· | ···················· | ····················· | ···················· | ···················· | ···················· |
| Z | 2336 | hCoV-19/Luxembourg/LNS1621763/2020\|EPI_ISL_739962\| | ···················· | ···················· | ····················· | ···················· | ······T············· | ···················· |
| Z | 2337 | hCoV-19/Luxembourg/LNS1632562/2020\|EPI_ISL_739963\| | ···················· | ···················· | ····················· | ···················· | ···················· | ···················· |
| Z | 2338 | hCoV-19/Luxembourg/LNS1634199/2020\|EPI_ISL_739964\| | ···················· | ···················· | ····················· | ···················· | ···················· | ···················· |
| Z | 2339 | hCoV-19/Luxembourg/LNS1636332/2020\|EPI_ISL_739965\| | ···················· | ···················· | ····················· | ···················· | ···················· | ···················· |
| Z | 2340 | hCoV-19/Luxembourg/LNS1639067/2020\|EPI_ISL_739966\| | ···················· | ···················· | ····················· | ···················· | ···················· | ···················· |
| Z | 2341 | hCoV-19/Luxembourg/LNS1643579/2020\|EPI_ISL_739967\| | ···················· | ···················· | ····················· | ···················· | ···················· | ···················· |
| Z | 2342 | hCoV-19/Luxembourg/LNS1644101/2020\|EPI_ISL_739968\| | ···················· | ···················· | ····················· | ···················· | ···················· | ···················· |
| Z | 2343 | hCoV-19/Luxembourg/LNS1651994/2020\|EPI_ISL_739969\| | ···················· | ···················· | ····················· | ···················· | ···················· | ···················· |
| Z | 2344 | hCoV-19/Luxembourg/LNS1664237/2020\|EPI_ISL_739970\| | ···················· | G··················· | ····················· | ···················· | ···················· | ···················· |
| Z | 2345 | hCoV-19/Luxembourg/LNS1665793/2020\|EPI_ISL_739971\| | ···················· | ···················· | ····················· | ···················· | ···················· | ···················· |
| Z | 2346 | hCoV-19/Luxembourg/LNS1669659/2020\|EPI_ISL_739972\| | ···················· | ···················· | ····················· | ···················· | ···················· | ···················· |
| Z | 2347 | hCoV-19/Luxembourg/LNS1670796/2020\|EPI_ISL_739973\| | ···················· | ···················· | ····················· | ···················· | ···················· | ···················· |
| Z | 2348 | hCoV-19/Luxembourg/LNS1673164/2020\|EPI_ISL_739974\| | ···················· | ···················· | ····················· | ···················· | ···················· | ···················· |
| Z | 2349 | hCoV-19/Luxembourg/LNS1682621/2020\|EPI_ISL_739975\| | ···················· | ···················· | ····················· | ···················· | ···················· | ···················· |
| Z | 2350 | hCoV-19/Luxembourg/LNS1686784/2020\|EPI_ISL_739976\| | ···················· | ···················· | ····················· | ···················· | ······T············· | ···················· |
| Z | 2351 | hCoV-19/Luxembourg/LNS1689160/2020\|EPI_ISL_739978\| | ···················· | ···················· | ····················· | ···················· | ···················· | ···················· |
| Z | 2352 | hCoV-19/Luxembourg/LNS1689912/2020\|EPI_ISL_739979\| | ···················· | ···················· | ····················· | ···················· | ···················· | ···················· |
| Z | 2353 | hCoV-19/Luxembourg/LNS1695013/2020\|EPI_ISL_739980\| | ···················· | ··········T········· | ····················· | ···················· | ···················· | ···················· |
| Z | 2354 | hCoV-19/Luxembourg/LNS1705026/2020\|EPI_ISL_739981\| | ···················· | ···················· | ····················· | ···················· | ···················· | ···················· |
| Z | 2355 | hCoV-19/Luxembourg/LNS1724590/2020\|EPI_ISL_739982\| | ···················· | G··················· | ····················· | ···················· | ···················· | ···················· |
| Z | 2356 | hCoV-19/Luxembourg/LNS1726450/2020\|EPI_ISL_739983\| | ···················· | ···················· | ····················· | ···················· | ···················· | ···················· |
| Z | 2357 | hCoV-19/Luxembourg/LNS1726686/2020\|EPI_ISL_739984\| | ···················· | ···················· | ····················· | ···················· | ···················· | ···················· |
| Z | 2358 | hCoV-19/Luxembourg/LNS1728273/2020\|EPI_ISL_739985\| | ···················· | ···················· | ····················· | ···················· | ···················· | ···················· |
| Z | 2359 | hCoV-19/Luxembourg/LNS1741668/2020\|EPI_ISL_739986\| | ···················· | ···················· | ····················· | ···················· | ···················· | ···················· |
| Z | 2360 | hCoV-19/Luxembourg/LNS1748642/2020\|EPI_ISL_739987\| | ···················· | ···················· | ····················· | ···················· | ···················· | ···················· |
| Z | 2361 | hCoV-19/Luxembourg/LNS1763709/2020\|EPI_ISL_739989\| | ···················· | ···················· | ····················· | ···················· | ···················· | ···················· |
| Z | 2362 | hCoV-19/Luxembourg/LNS1782150/2020\|EPI_ISL_739990\| | ···················· | ···················· | ····················· | ···················· | ···················· | ···················· |
| Z | 2363 | hCoV-19/Luxembourg/LNS1785999/2020\|EPI_ISL_739991\| | ···················· | ···················· | ····················· | ···················· | ···················· | ···················· |
| Z | 2364 | hCoV-19/Luxembourg/LNS1798053/2020\|EPI_ISL_739993\| | ···················· | ···················· | ····················· | ···················· | ···················· | ···················· |
| Z | 2365 | hCoV-19/Luxembourg/LNS1807556/2020\|EPI_ISL_739995\| | ···················· | ···················· | ····················· | ···················· | ···················· | ···················· |
| Z | 2366 | hCoV-19/Luxembourg/LNS1807965/2020\|EPI_ISL_739996\| | ···················· | ···················· | ····················· | ···················· | ···················· | ···················· |
| Z | 2367 | hCoV-19/Luxembourg/LNS1819970/2020\|EPI_ISL_739997\| | ···················· | ···················· | ····················· | ···················· | ···················· | ···················· |
| Z | 2368 | hCoV-19/Luxembourg/LNS1825574/2020\|EPI_ISL_739998\| | ···················· | ···················· | ····················· | ···················· | ···················· | ···················· |
| Z | 2369 | hCoV-19/Luxembourg/LNS1827474/2020\|EPI_ISL_739999\| | ···················· | ···················· | ····················· | ···················· | ···················· | ···················· |
| Z | 2370 | hCoV-19/England/ALDP-B75205/2020\|EPI_ISL_740900\|20 | ···················· | ···················· | ····················· | ···················· | ···················· | ···················· |
| Z | 2371 | hCoV-19/England/ALDP-B75223/2020\|EPI_ISL_740901\|20 | ···················· | ···················· | ····················· | ···················· | ···················· | ···················· |
| Z | 2372 | hCoV-19/England/ALDP-B75232/2020\|EPI_ISL_740902\|20 | ···················· | ···················· | ····················· | ···················· | ···················· | ···················· |
| Z | 2373 | hCoV-19/England/ALDP-B75241/2020\|EPI_ISL_740903\|20 | ···················· | ···················· | ····················· | ···················· | ···················· | ···················· |
| Z | 2374 | hCoV-19/England/ALDP-B75469/2020\|EPI_ISL_740904\|20 | ···················· | ···················· | ····················· | ···················· | ···················· | ···················· |
| Z | 2375 | hCoV-19/England/ALDP-B754A5/2020\|EPI_ISL_740906\|20 | ···················· | ···················· | ····················· | ···················· | ···················· | ···················· |
| Z | 2376 | hCoV-19/England/ALDP-B754B4/2020\|EPI_ISL_740907\|20 | ···················· | ···················· | ····················· | ···················· | ···················· | ···················· |
| Z | 2377 | hCoV-19/England/ALDP-B754D2/2020\|EPI_ISL_740908\|20 | ···················· | ···················· | ····················· | ···················· | ···················· | ···················· |
| Z | 2378 | hCoV-19/England/ALDP-B754E1/2020\|EPI_ISL_740909\|20 | ···················· | ···················· | ····················· | ···················· | ···················· | ···················· |
| Z | 2379 | hCoV-19/England/ALDP-B7551B/2020\|EPI_ISL_740910\|20 | ···················· | ···················· | ····················· | ···················· | ···················· | ···················· |
| Z | 2380 | hCoV-19/England/ALDP-B7552A/2020\|EPI_ISL_740911\|20 | ···················· | ···················· | ····················· | ···················· | ···················· | ···················· |
| Z | 2381 | hCoV-19/England/ALDP-B75539/2020\|EPI_ISL_740912\|20 | ···················· | ···················· | ····················· | ···················· | ···················· | ···················· |
| Z | 2382 | hCoV-19/England/ALDP-B75663/2020\|EPI_ISL_740913\|20 | ···················· | ···················· | ····················· | ···················· | ···················· | ···················· |
| Z | 2383 | hCoV-19/England/ALDP-B75681/2020\|EPI_ISL_740914\|20 | ···················· | ···················· | ····················· | ···················· | ···················· | ···················· |
| Z | 2384 | hCoV-19/England/ALDP-B75690/2020\|EPI_ISL_740915\|20 | ···················· | ···················· | ····················· | ···················· | ···················· | ···················· |
| Z | 2385 | hCoV-19/England/ALDP-B756AF/2020\|EPI_ISL_740916\|20 | ···················· | ···················· | ····················· | ···················· | ···················· | ···················· |
| Z | 2386 | hCoV-19/England/ALDP-B756BE/2020\|EPI_ISL_740917\|20 | ···················· | ···················· | ····················· | ···················· | ···················· | ···················· |
| Z | 2387 | hCoV-19/England/ALDP-B756DC/2020\|EPI_ISL_740918\|20 | ···················· | ···················· | ····················· | ···················· | ···················· | ···················· |
| Z | 2388 | hCoV-19/England/ALDP-B75715/2020\|EPI_ISL_740919\|20 | ···················· | ···················· | ····················· | ···················· | ···················· | ···················· |
| Z | 2389 | hCoV-19/England/ALDP-B75724/2020\|EPI_ISL_740920\|20 | ···················· | ·A·················· | ····················· | ···················· | ···················· | ···················· |
| Z | 2390 | hCoV-19/England/ALDP-B75760/2020\|EPI_ISL_740921\|20 | ···················· | ···················· | ····················· | ···················· | ···················· | ···················· |
| Z | 2391 | hCoV-19/England/ALDP-B7578E/2020\|EPI_ISL_740922\|20 | ···················· | ···················· | ····················· | ···················· | ···················· | ···················· |
| Z | 2392 | hCoV-19/England/ALDP-B757AC/2020\|EPI_ISL_740923\|20 | ···················· | ·A·················· | ····················· | ···················· | ···················· | ···················· |
| Z | 2393 | hCoV-19/England/ALDP-B757F7/2020\|EPI_ISL_740924\|20 | ···················· | ···················· | ····················· | ···················· | ···················· | ···················· |

| | | | | | | | | |
|---|---|---|---|---|---|---|---|---|
| Z | 2394 | hCoV-19/England/ALDP-B75803/2020\|EPI_ISL_740925\|20 | ···················· | ···················· | ···················· | ···················· | ···················· | ···················· |
| Z | 2395 | hCoV-19/England/ALDP-B7595B/2020\|EPI_ISL_740928\|20 | ···················· | ···················· | ···················· | ···················· | ···················· | ···················· |
| Z | 2396 | hCoV-19/England/ALDP-B75979/2020\|EPI_ISL_740929\|20 | ···················· | ···················· | ···················· | ···················· | ···················· | ···················· |
| Z | 2397 | hCoV-19/England/ALDP-B75B91/2020\|EPI_ISL_740940\|20 | ···················· | ···················· | ···················· | ···················· | ···················· | ···················· |
| Z | 2398 | hCoV-19/England/ALDP-B75BCE/2020\|EPI_ISL_740941\|20 | ···················· | ···················· | ···················· | ···················· | ···················· | ···················· |
| Z | 2399 | hCoV-19/England/ALDP-B75BEC/2020\|EPI_ISL_740943\|20 | ···················· | ···················· | ···················· | ···················· | ···················· | ···················· |
| Z | 2400 | hCoV-19/England/ALDP-B75C16/2020\|EPI_ISL_740944\|20 | ···················· | ···················· | ···················· | ···················· | ···················· | ···················· |
| Z | 2401 | hCoV-19/England/ALDP-B75E98/2020\|EPI_ISL_740945\|20 | ···················· | ···················· | ···················· | ···················· | ···················· | ···················· |
| Z | 2402 | hCoV-19/England/ALDP-B75EA7/2020\|EPI_ISL_740946\|20 | ···················· | ···················· | ···················· | ···················· | ···················· | ···················· |
| Z | 2403 | hCoV-19/England/ALDP-B75EB6/2020\|EPI_ISL_740947\|20 | ···················· | ···················· | ···················· | ···················· | ···················· | ···················· |
| Z | 2404 | hCoV-19/England/ALDP-B75EC5/2020\|EPI_ISL_740948\|20 | ···················· | ···················· | ···················· | ···················· | ···················· | ···················· |
| Z | 2405 | hCoV-19/England/ALDP-B75ED4/2020\|EPI_ISL_740949\|20 | ···················· | ···················· | ···················· | ···················· | ···················· | ···················· |
| Z | 2406 | hCoV-19/England/ALDP-B75EF2/2020\|EPI_ISL_740950\|20 | ···················· | ···················· | ···················· | ···················· | ···················· | ···················· |
| Z | 2407 | hCoV-19/England/ALDP-B75F3B/2020\|EPI_ISL_740951\|20 | ···················· | ···················· | ···················· | ···················· | ···················· | ···················· |
| Z | 2408 | hCoV-19/England/ALDP-B75F68/2020\|EPI_ISL_740952\|20 | ···················· | ···················· | ···················· | ···················· | ···················· | ···················· |
| Z | 2409 | hCoV-19/England/CAMB-1BC4CA/2020\|EPI_ISL_740954\|20 | ···················· | ···················· | ···················· | ···················· | ···················· | ···················· |
| Z | 2410 | hCoV-19/England/CAMB-1BCCFF/2020\|EPI_ISL_740955\|20 | ···················· | ···················· | ···················· | ···················· | ···················· | ···················· |
| Z | 2411 | hCoV-19/England/CAMB-1BD04D/2020\|EPI_ISL_740957\|20 | ···················· | ···················· | ···················· | ···················· | ···················· | ···················· |
| Z | 2412 | hCoV-19/England/CAMB-1BD687/2020\|EPI_ISL_740968\|20 | ···················· | ···················· | ···················· | ···················· | ···················· | ···················· |
| Z | 2413 | hCoV-19/England/CAMB-1BD696/2020\|EPI_ISL_740969\|20 | ···················· | ···················· | ···················· | ···················· | ···················· | ···················· |
| Z | 2414 | hCoV-19/England/CAMB-1BD6C3/2020\|EPI_ISL_740972\|20 | ···················· | ···················· | ···················· | ···················· | ···················· | ···················· |
| Z | 2415 | hCoV-19/England/CAMB-1BD739/2020\|EPI_ISL_740977\|20 | ···················· | ···················· | ···················· | ···················· | ···················· | ···················· |
| Z | 2416 | hCoV-19/England/CAMB-1BD748/2020\|EPI_ISL_740978\|20 | ···················· | ···················· | ···················· | ···················· | ···················· | ···················· |
| Z | 2417 | hCoV-19/England/CAMB-1BD766/2020\|EPI_ISL_740979\|20 | ···················· | ···················· | ···················· | ···················· | ···················· | ···················· |
| Z | 2418 | hCoV-19/England/CAMB-1BD784/2020\|EPI_ISL_740980\|20 | ···················· | ···················· | ···················· | ···················· | ···················· | ···················· |
| Z | 2419 | hCoV-19/England/CAMB-1BD7B1/2020\|EPI_ISL_740981\|20 | ···················· | ···················· | ···················· | ···················· | ···················· | ···················· |
| Z | 2420 | hCoV-19/England/CAMB-1BD818/2020\|EPI_ISL_740984\|20 | ···················· | ·A·················· | ···················· | ···················· | ···················· | ···A················ |
| Z | 2421 | hCoV-19/England/CAMB-1BD8CD/2020\|EPI_ISL_740987\|20 | ···················· | ···················· | ···················· | ···················· | ···················· | ···················· |
| Z | 2422 | hCoV-19/England/CAMB-1BD8DC/2020\|EPI_ISL_740988\|20 | ···················· | ···················· | ···················· | ···················· | ···················· | ···················· |
| Z | 2423 | hCoV-19/England/CAMB-1BD906/2020\|EPI_ISL_740989\|20 | ···················· | ···················· | ···················· | ···················· | ···················· | ···················· |
| Z | 2428 | hCoV-19/Wales/PHWC-49F266/2020\|EPI_ISL_743186\|2020 | ···················· | ···················· | ···················· | ···················· | ···················· | ···················· |
| Z | 2430 | hCoV-19/Wales/PHWC-49F293/2020\|EPI_ISL_743189\|2020 | ···················· | ······A············· | ···················· | ···················· | ···················· | ···················· |
| Z | 2431 | hCoV-19/Wales/PHWC-49F2A2/2020\|EPI_ISL_743190\|2020 | ···················· | ···················· | ···················· | ···················· | ···················· | ···················· |
| Z | 2432 | hCoV-19/Wales/PHWC-49F2B1/2020\|EPI_ISL_743191\|2020 | ···················· | ···················· | ···················· | ···················· | ···················· | ···················· |
| Z | 2433 | hCoV-19/Wales/PHWC-49F2DF/2020\|EPI_ISL_743193\|2020 | ···················· | ···················· | ···················· | ···················· | ···················· | ···················· |
| Z | 2436 | hCoV-19/Wales/PHWC-49F327/2020\|EPI_ISL_743198\|2020 | ···················· | ···················· | ···················· | ···················· | ···················· | ···················· |
| Z | 2438 | hCoV-19/Wales/PHWC-49F363/2020\|EPI_ISL_743202\|2020 | ···················· | ···················· | ···················· | ···················· | ···················· | ···················· |
| Z | 2440 | hCoV-19/Wales/PHWC-49F381/2020\|EPI_ISL_743204\|2020 | ···················· | ···················· | ···················· | ···················· | ···················· | ···················· |
| Z | 2441 | hCoV-19/Wales/PHWC-49F390/2020\|EPI_ISL_743205\|2020 | ···················· | ···················· | ···················· | ···················· | ···················· | ···················· |
| Z | 2442 | hCoV-19/Wales/PHWC-49F3AF/2020\|EPI_ISL_743206\|2020 | ···················· | ···················· | ···················· | ···················· | ···················· | ···················· |
| Z | 2443 | hCoV-19/Wales/PHWC-49F3BE/2020\|EPI_ISL_743207\|2020 | ···················· | ···················· | ···················· | ···················· | ···················· | ···················· |
| Z | 2445 | hCoV-19/Wales/PHWC-49F3DC/2020\|EPI_ISL_743209\|2020 | ···················· | ······A············· | ···················· | ···················· | ···················· | ···················· |
| Z | 2446 | hCoV-19/Wales/PHWC-49F3EB/2020\|EPI_ISL_743210\|2020 | ···················· | ···················· | ···················· | ···················· | ···················· | ···················· |
| Z | 2447 | hCoV-19/Wales/PHWC-49F406/2020\|EPI_ISL_743212\|2020 | ···················· | ···················· | ···················· | ···················· | ···················· | ···················· |
| Z | 2448 | hCoV-19/Wales/PHWC-49F415/2020\|EPI_ISL_743213\|2020 | ···················· | ···················· | ···················· | ···················· | ···················· | ···················· |
| Z | 2449 | hCoV-19/Wales/PHWC-49F442/2020\|EPI_ISL_743216\|2020 | ···················· | ···················· | ···················· | ···················· | ···················· | ···················· |
| Z | 2450 | hCoV-19/Luxembourg/LNS7041741/2020\|EPI_ISL_744500\| | ···················· | ···················· | ···················· | ···················· | ···················· | ···················· |
| Z | 2451 | hCoV-19/Luxembourg/LNS7042092/2020\|EPI_ISL_744501\| | ···················· | ···················· | ···················· | ···················· | ···················· | ···················· |
| Z | 2452 | hCoV-19/Luxembourg/LNS7047940/2020\|EPI_ISL_744502\| | ···················· | ···················· | ···················· | ···················· | ···················· | ···················· |
| Z | 2453 | hCoV-19/Luxembourg/LNS7062667/2020\|EPI_ISL_744503\| | ···················· | ···················· | ···················· | ···················· | ···················· | ···················· |
| Z | 2454 | hCoV-19/Luxembourg/LNS7088375/2020\|EPI_ISL_744510\| | ···················· | ···················· | ···················· | ················T··· | ···················· | ···················· |
| Z | 2455 | hCoV-19/Luxembourg/LNS7093842/2020\|EPI_ISL_744511\| | ···················· | ···················· | ···················· | ···················· | ···················· | ···················· |
| Z | 2456 | hCoV-19/Luxembourg/LNS7100858/2020\|EPI_ISL_744512\| | ···················· | ···················· | ···················· | ···················· | ···················· | ···················· |
| Z | 2457 | hCoV-19/Luxembourg/LNS7106723/2020\|EPI_ISL_744513\| | ···················· | ···················· | ···················· | ···················· | ···················· | ···················· |
| Z | 2458 | hCoV-19/Luxembourg/LNS7106836/2020\|EPI_ISL_744514\| | ···················· | ···················· | ···················· | ···················· | ···················· | ···················· |
| Z | 2459 | hCoV-19/Luxembourg/LNS7114654/2020\|EPI_ISL_744515\| | ···················· | ···················· | ···················· | ···················· | ···················· | ···················· |
| Z | 2460 | hCoV-19/Luxembourg/LNS7115373/2020\|EPI_ISL_744516\| | ···················· | ···················· | ···················· | ···················· | ···················· | ···················· |
| Z | 2461 | hCoV-19/Luxembourg/LNS7117267/2020\|EPI_ISL_744517\| | ···················· | ···················· | ···················· | ···················· | ···················· | ···················· |
| Z | 2462 | hCoV-19/Luxembourg/LNS7121709/2020\|EPI_ISL_744518\| | ···················· | ···················· | ···················· | ···················· | ···················· | ···················· |
| Z | 2463 | hCoV-19/Luxembourg/LNS7124526/2020\|EPI_ISL_744519\| | ···················· | ···················· | ···········T········ | ···················· | ···················· | ···················· |
| Z | 2464 | hCoV-19/Luxembourg/LNS7132932/2020\|EPI_ISL_744521\| | ············T······· | ···················· | ···················· | ···················· | ···················· | ···················· |
| Z | 2465 | hCoV-19/Luxembourg/LNS7147643/2020\|EPI_ISL_744522\| | ···················· | ··········T········· | ···················· | ···················· | ···················· | ···················· |
| Z | 2466 | hCoV-19/Luxembourg/LNS7158995/2020\|EPI_ISL_744524\| | ···················· | ···················· | ···················· | ···················· | ···················· | ···················· |
| Z | 2467 | hCoV-19/Luxembourg/LNS7174945/2020\|EPI_ISL_744525\| | ···················· | G··················· | ···················· | ···················· | ···················· | ···················· |
| Z | 2468 | hCoV-19/Luxembourg/LNS7175419/2020\|EPI_ISL_744526\| | ···················· | ···················· | ···················· | ···················· | ···················· | ···················· |
| Z | 2469 | hCoV-19/Luxembourg/LNS7181694/2020\|EPI_ISL_744527\| | ···················· | ···················· | ···················· | ···················· | ···················· | ···················· |
| Z | 2470 | hCoV-19/Luxembourg/LNS7184421/2020\|EPI_ISL_744528\| | ···················· | ···················· | ···················· | ···················· | ···················· | ···················· |
| Z | 2471 | hCoV-19/Luxembourg/LNS7186551/2020\|EPI_ISL_744529\| | ···················· | ···················· | ···················· | ···················· | ···················· | ···················· |
| Z | 2472 | hCoV-19/Luxembourg/LNS7194612/2020\|EPI_ISL_744530\| | ···················· | ···················· | ···················· | ···················· | ···················· | ···················· |
| Z | 2473 | hCoV-19/Luxembourg/LNS7199551/2020\|EPI_ISL_744533\| | ···················· | ···················· | ···················· | ···················· | ···················· | ···················· |
| Z | 2474 | hCoV-19/Luxembourg/LNS7201440/2020\|EPI_ISL_744534\| | ···················· | ···················· | ···················· | ···················· | ···················· | ···················· |
| Z | 2475 | hCoV-19/Luxembourg/LNS7202999/2020\|EPI_ISL_744535\| | ···················· | ···················· | ···················· | ···················· | ···················· | ···················· |
| Z | 2476 | hCoV-19/Luxembourg/LNS7205423/2020\|EPI_ISL_744536\| | ···················· | ···················· | ···················· | ···················· | ···················· | ···················· |
| Z | 2477 | hCoV-19/Luxembourg/LNS7215500/2020\|EPI_ISL_744537\| | ···················· | ···················· | ···················· | ···················· | ···················· | ···················· |
| Z | 2478 | hCoV-19/Luxembourg/LNS7224825/2020\|EPI_ISL_744538\| | ···················· | ···················· | ···················· | ···················· | ···················· | ···················· |
| Z | 2479 | hCoV-19/Luxembourg/LNS7228271/2020\|EPI_ISL_744539\| | ···················· | ···················· | ···················· | ···················· | ···················· | ···················· |
| Z | 2480 | hCoV-19/Luxembourg/LNS7233124/2020\|EPI_ISL_744540\| | ···················· | ···················· | ···················· | ···················· | ···················· | ···················· |
| Z | 2481 | hCoV-19/Luxembourg/LNS7238398/2020\|EPI_ISL_744541\| | ···················· | ···················· | ···················· | ···················· | ···················· | ···················· |
| Z | 2482 | hCoV-19/Luxembourg/LNS7243430/2020\|EPI_ISL_744542\| | ···················· | ···················· | ···················· | ···················· | ···················· | ···················· |
| Z | 2483 | hCoV-19/Luxembourg/LNS7259264/2020\|EPI_ISL_744543\| | ···················· | ···················· | ···················· | ···················· | ···················· | ···················· |

Z 2484 hCoV-19/Luxembourg/LNS7261025/2020|EPI_ISL_744544| ···················· ···················· ···················· ···················· ···················· ····················
Z 2485 hCoV-19/Luxembourg/LNS7278260/2020|EPI_ISL_744545| ···················· ···················· ···················· ···················· ···················· ····················
Z 2486 hCoV-19/Luxembourg/LNS7292249/2020|EPI_ISL_744546| ···················· ···················· ···················· ···················· ···················· ····················
Z 2487 hCoV-19/Luxembourg/LNS7293580/2020|EPI_ISL_744547| ···················· ···················· ···················· ···················· ···················· ····················
Z 2488 hCoV-19/USA/UT-UPHL-2012190289/2020|EPI_ISL_745800 ···················· ···················· ···················· ···················· ···················· ····················
Z 2489 hCoV-19/USA/UT-UPHL-2012736957/2020|EPI_ISL_745801 ···················· ···················· ···················· ···················· ···················· ····················
Z 2490 hCoV-19/USA/UT-UPHL-2012942531/2020|EPI_ISL_745802 ···················· ···················· ···················· ···················· ···················· ····················
Z 2491 hCoV-19/USA/UT-UPHL-2012842708/2020|EPI_ISL_745803 ···················· ···················· ···················· ···················· ···················· ····················
Z 2492 hCoV-19/USA/UT-UPHL-2012275073/2020|EPI_ISL_745804 ···················· ···················· ···················· ···················· ···················· ····················
Z 2493 hCoV-19/USA/UT-UPHL-2012706876/2020|EPI_ISL_745805 ···················· ···················· ···················· ···················· ···················· ····················
Z 2494 hCoV-19/USA/UT-UPHL-2012606373/2020|EPI_ISL_745806 ···················· ···················· ···················· ···················· ···················· ····················
Z 2495 hCoV-19/USA/UT-UPHL-2012818468/2020|EPI_ISL_745807 ···················· ···················· ···················· ···················· ···················· ····················
Z 2496 hCoV-19/USA/UT-UPHL-2012770703/2020|EPI_ISL_745808 ···················· ···················· ···················· ···················· ···················· ····················
Z 2497 hCoV-19/USA/UT-UPHL-2012189523/2020|EPI_ISL_745809 ···················· ···················· ···················· ···················· ···················· ····················
Z 2498 hCoV-19/USA/UT-UPHL-2012274873/2020|EPI_ISL_745810 ···················· ···················· ···················· ···················· ···················· ····················
Z 2499 hCoV-19/USA/UT-UPHL-2012905899/2020|EPI_ISL_745811 ···················· ···················· ···················· ···················· ···················· ····················
Z 2500 hCoV-19/USA/UT-UPHL-2012436073/2020|EPI_ISL_745812 ···················· ···················· ···················· ···················· ···················· ····················
Z 2501 hCoV-19/USA/UT-UPHL-2012502791/2020|EPI_ISL_745813 ···················· ···················· ···················· ···················· ···················· ····················
Z 2502 hCoV-19/USA/UT-UPHL-2012321043/2020|EPI_ISL_745814 ···················· ···················· ···················· ···················· ···················· ····················
Z 2503 hCoV-19/USA/UT-UPHL-2012082456/2020|EPI_ISL_745815 ···················· ···················· ···················· ···················· ···················· ····················
Z 2504 hCoV-19/USA/UT-UPHL-2012436214/2020|EPI_ISL_745816 ···················· ···················· ···················· ···················· ···················· ····················
Z 2505 hCoV-19/USA/UT-UPHL-2012812052/2020|EPI_ISL_745817 ···················· ···················· ···················· ···················· ···················· ····················
Z 2506 hCoV-19/USA/UT-UPHL-2012069108/2020|EPI_ISL_745818 ···················· ·A·················· ···················· ···················· ···················· ····················
Z 2507 hCoV-19/USA/UT-UPHL-2012434246/2020|EPI_ISL_745819 ···················· ···················· ···················· ···················· ···················· ····················
Z 2508 hCoV-19/USA/UT-UPHL-2012154383/2020|EPI_ISL_745820 ···················· ···················· ···················· ···················· ···················· ····················
Z 2509 hCoV-19/USA/UT-UPHL-2012332038/2020|EPI_ISL_745821 ···················· ···················· ···················· ···················· ···················· ····················
Z 2510 hCoV-19/USA/UT-UPHL-2012742622/2020|EPI_ISL_745822 ···················· ···················· ···················· ···················· ···················· ····················
Z 2511 hCoV-19/USA/UT-UPHL-2012886729/2020|EPI_ISL_745823 ···················· ···················· ···················· ···················· ···················· ····················
Z 2512 hCoV-19/USA/UT-UPHL-2012156788/2020|EPI_ISL_745824 ···················· ···················· ···················· ···················· ···················· ····················
Z 2513 hCoV-19/USA/UT-UPHL-2012789048/2020|EPI_ISL_745825 ···················· ···················· ···················· ···················· ···················· ····················
Z 2514 hCoV-19/USA/UT-UPHL-2012280782/2020|EPI_ISL_745826 ···················· ·A·················· ···················· ···················· ···················· ····················
Z 2515 hCoV-19/USA/UT-UPHL-2012538042/2020|EPI_ISL_745827 ···················· ···················· ···················· ···················· ···················· ····················
Z 2516 hCoV-19/USA/UT-UPHL-2012090434/2020|EPI_ISL_745828 ···················· ···················· ···················· ···················· ···················· ····················
Z 2517 hCoV-19/USA/UT-UPHL-2012208753/2020|EPI_ISL_745829 ···················· ·A·················· ···················· ···················· ···················· ····················
Z 2518 hCoV-19/USA/UT-UPHL-2012581328/2020|EPI_ISL_745830 ···················· ·A·················· ···················· ···················· ···················· ····················
Z 2519 hCoV-19/USA/UT-UPHL-2012145663/2020|EPI_ISL_745831 ···················· ···················· ···················· ···················· ···················· ····················
Z 2520 hCoV-19/USA/UT-UPHL-2012063197/2020|EPI_ISL_745832 ············T······· ···················· ···················· ···················· ···················· ····················
Z 2521 hCoV-19/USA/UT-UPHL-2012080415/2020|EPI_ISL_745833 ···················· ·A·················· ···················· ···················· ···················· ····················
Z 2522 hCoV-19/USA/UT-UPHL-2012487788/2020|EPI_ISL_745834 ···················· ·A·················· ···················· ···················· ···················· ····················
Z 2523 hCoV-19/USA/UT-UPHL-2012346545/2020|EPI_ISL_745840 ···················· ·A·················· ···················· ···················· ···················· ····················
Z 2524 hCoV-19/USA/UT-UPHL-2012250967/2020|EPI_ISL_745841 ···················· ···················· ···················· ···················· ···················· ····················
Z 2525 hCoV-19/USA/UT-UPHL-2012719632/2020|EPI_ISL_745842 ···················· ···················· ···················· ···················· ···················· ····················
Z 2526 hCoV-19/USA/UT-UPHL-2012138984/2020|EPI_ISL_745844 ···················· ···················· ···················· ···················· ···················· ····················
Z 2527 hCoV-19/USA/UT-UPHL-2012380154/2020|EPI_ISL_745845 ···················· ···················· ···················· ···················· ···················· ····················
Z 2528 hCoV-19/USA/UT-UPHL-2012526711/2020|EPI_ISL_745846 ···················· ···················· ···················· ···················· ···················· ····················
Z 2529 hCoV-19/USA/UT-UPHL-2012383134/2020|EPI_ISL_745847 ···················· ···················· ···················· ···················· ···················· ····················
Z 2530 hCoV-19/USA/UT-UPHL-2012753144/2020|EPI_ISL_745848 ···················· ···················· ···················· ···················· ···················· ····················
Z 2531 hCoV-19/USA/UT-UPHL-2012943778/2020|EPI_ISL_745849 ···················· ···················· ···················· ···················· ···················· ····················
Z 2532 hCoV-19/USA/UT-UPHL-2012671868/2020|EPI_ISL_745850 ···················· ·A·················· ···················· ···················· ···················· ····················
Z 2533 hCoV-19/USA/UT-UPHL-2012842916/2020|EPI_ISL_745851 ···················· ···················· ···················· ···················· ···················· ····················
Z 2534 hCoV-19/USA/UT-UPHL-2012353321/2020|EPI_ISL_745852 ···················· ···················· ···················· ···················· ···················· ····················
Z 2535 hCoV-19/USA/UT-UPHL-2012571363/2020|EPI_ISL_745853 ···················· ·A·················· ···················· ···················· ···················· ····················
Z 2536 hCoV-19/USA/UT-UPHL-2012584910/2020|EPI_ISL_745854 ···················· ···················· ···················· ···················· ···················· ····················
Z 2537 hCoV-19/USA/UT-UPHL-2012766698/2020|EPI_ISL_745855 ···················· ···················· ···················· ···················· ···················· ····················
Z 2538 hCoV-19/USA/UT-UPHL-2012074313/2020|EPI_ISL_745856 ···················· ···················· ···················· ···················· ···················· ····················
Z 2539 hCoV-19/USA/UT-UPHL-2012846734/2020|EPI_ISL_745857 ···················· ·A·················· ···················· ···················· ···················· ····················
Z 2540 hCoV-19/USA/UT-UPHL-2012084152/2020|EPI_ISL_745858 ···················· ·A·················· ···················· ···················· ···················· ····················
Z 2541 hCoV-19/USA/UT-UPHL-2012191494/2020|EPI_ISL_745859 ···················· ·A·················· ···················· ···················· ···················· ····················
Z 2542 hCoV-19/USA/UT-UPHL-2012214057/2020|EPI_ISL_745860 ···················· ···················· ···················· ···················· ···················· ····················
Z 2543 hCoV-19/USA/UT-UPHL-2012514756/2020|EPI_ISL_745861 ···················· ·A·················· ···················· ···················· ···················· ····················
Z 2544 hCoV-19/USA/UT-UPHL-2012020588/2020|EPI_ISL_745862 ···················· ···················· ···················· ···················· ···················· ····················
Z 2545 hCoV-19/USA/UT-UPHL-2012724093/2020|EPI_ISL_745863 ···················· ···················· ···················· ···················· ···················· ····················
Z 2546 hCoV-19/USA/UT-UPHL-2012401286/2020|EPI_ISL_745864 ···················· ···················· ···················· ···················· ···················· ····················
Z 2547 hCoV-19/USA/UT-UPHL-2012621057/2020|EPI_ISL_745865 ···················· ···················· ···················· ···················· ···················· ····················
Z 2548 hCoV-19/USA/UT-UPHL-2012306516/2020|EPI_ISL_745866 ···················· ···················· ···················· ···················· ···················· ····················
Z 2549 hCoV-19/USA/UT-UPHL-2012160224/2020|EPI_ISL_745867 ···················· ···················· ···················· ···················· ···················· ····················
Z 2550 hCoV-19/USA/UT-UPHL-2012803399/2020|EPI_ISL_745868 ···················· ···················· ···················· ···················· ···················· ····················
Z 2551 hCoV-19/USA/UT-UPHL-2012766678/2020|EPI_ISL_745869 ···················· ···················· ···················· ···················· ···················· ····················
Z 2552 hCoV-19/USA/UT-UPHL-2012818011/2020|EPI_ISL_745870 ···················· ···················· ···················· ···················· ···················· ····················
Z 2553 hCoV-19/USA/UT-UPHL-2012046308/2020|EPI_ISL_745871 ···················· ·A·················· ···················· ···················· ···················· ····················
Z 2554 hCoV-19/USA/UT-UPHL-2012685247/2020|EPI_ISL_745872 ···················· ···················· ···················· ···················· ···················· ····················
Z 2555 hCoV-19/USA/UT-UPHL-2012810942/2020|EPI_ISL_745873 ···················· ···················· ···················· ···················· ···················· ····················
Z 2556 hCoV-19/USA/UT-UPHL-2012476056/2020|EPI_ISL_745874 ···················· ···················· ···················· ···················· ···················· ····················
Z 2557 hCoV-19/USA/UT-UPHL-2012905638/2020|EPI_ISL_745875 ···················· ···················· ···················· ···················· ···················· ····················
Z 2558 hCoV-19/USA/UT-UPHL-2012155157/2020|EPI_ISL_745876 ···················· ···················· ···················· ···················· ···················· ····················
Z 2559 hCoV-19/USA/UT-UPHL-2012890011/2020|EPI_ISL_745877 ···················· ···················· ···················· ···················· ···················· ····················
Z 2560 hCoV-19/USA/UT-UPHL-2012370082/2020|EPI_ISL_745878 ···················· ·A·················· ···················· ···················· ···················· ····················
Z 2561 hCoV-19/USA/CA-CZB-14149/2020|EPI_ISL_738707|2020- ···················· ·A·················· ···················· ···················· ···················· ····················
Z 2562 hCoV-19/USA/CA-CZB-14148/2020|EPI_ISL_738708|2020- ···················· ·A·················· ···················· ···················· ···················· ····················
Z 2563 hCoV-19/USA/CA-CZB-14663/2020|EPI_ISL_738709|2020- ···················· ···················· ···················· ···················· ···················· ····················

| | | | | | | | | |
|---|---|---|---|---|---|---|---|---|
| Z | 2564 | hCoV-19/USA/CA-CZB-14962/2020\|EPI_ISL_738710\|2020- | ···················· | ·A·················· | ···················· | ···················· | ···················· | ···················· |
| Z | 2565 | hCoV-19/USA/CA-CZB-14247/2020\|EPI_ISL_738711\|2020- | ···················· | ···················· | ···················· | ···················· | ···················· | ···················· |
| Z | 2566 | hCoV-19/USA/CA-CZB-16183/2020\|EPI_ISL_738712\|2020- | ···················· | ···················· | ···················· | ···················· | ···················· | ···················· |
| Z | 2567 | hCoV-19/USA/CA-CZB-16170/2020\|EPI_ISL_738713\|2020- | ···················· | ···················· | ···················· | ···················· | ···················· | ···················· |
| Z | 2568 | hCoV-19/USA/CA-CZB-16171/2020\|EPI_ISL_738714\|2020- | ···················· | ···················· | ···················· | ···················· | ···················· | ···················· |
| Z | 2569 | hCoV-19/USA/CA-CZB-14414/2020\|EPI_ISL_738715\|2020- | ···················· | ···················· | ···················· | ···················· | ···················· | ···················· |
| Z | 2570 | hCoV-19/USA/CA-CZB-14672/2020\|EPI_ISL_738722\|2020- | ···················· | ···················· | ···················· | ···················· | ···················· | ···················· |
| Z | 2571 | hCoV-19/USA/CA-CZB-15987/2020\|EPI_ISL_738723\|2020- | ···················· | ···················· | ···················· | ···················· | ···················· | ···················· |
| Z | 2572 | hCoV-19/USA/CA-CZB-14128/2020\|EPI_ISL_738724\|2020- | ···················· | ···················· | ···················· | ···················· | ···················· | ···················· |
| Z | 2573 | hCoV-19/USA/CA-CZB-16145/2020\|EPI_ISL_738725\|2020- | ···················· | ···················· | ···················· | ···················· | ···················· | ···················· |
| Z | 2575 | hCoV-19/USA/CA-CZB-16172/2020\|EPI_ISL_738727\|2020- | ···················· | ···················· | ···················· | ···················· | ···················· | ···················· |
| Z | 2576 | hCoV-19/USA/CA-CZB-14541/2020\|EPI_ISL_738728\|2020- | ···················· | ·A·················· | ···················· | ···················· | ···················· | ···················· |
| Z | 2577 | hCoV-19/USA/CA-CZB-14677/2020\|EPI_ISL_738729\|2020- | ···················· | ···················· | ···················· | ···················· | ···················· | ···················· |
| Z | 2578 | hCoV-19/USA/CA-CZB-14539/2020\|EPI_ISL_738730\|2020- | ···················· | ·A·················· | ···················· | ···················· | ···················· | ···················· |
| Z | 2579 | hCoV-19/USA/CA-CZB-14770/2020\|EPI_ISL_738742\|2020- | ···················· | ·A·················· | ···················· | ···················· | ···················· | ···················· |
| Z | 2581 | hCoV-19/USA/CA-CZB-13816/2020\|EPI_ISL_738744\|2020- | ···················· | ···················· | ···················· | ···················· | ···················· | ···················· |
| Z | 2583 | hCoV-19/USA/CA-CZB-15072/2020\|EPI_ISL_738746\|2020- | ···················· | ···················· | ···················· | ···················· | ···················· | ···················· |
| Z | 2584 | hCoV-19/USA/CA-CZB-15341/2020\|EPI_ISL_738747\|2020- | ···················· | ···················· | ···················· | ···················· | ···················· | ···················· |
| Z | 2587 | hCoV-19/USA/CA-CZB-15979/2020\|EPI_ISL_738750\|2020- | ···················· | ···················· | ···················· | ···················· | ···················· | ···················· |
| Z | 2588 | hCoV-19/USA/CA-CZB-14012/2020\|EPI_ISL_738751\|2020- | ···················· | ···················· | ···················· | ···················· | ···················· | ···················· |
| Z | 2589 | hCoV-19/USA/CA-CZB-15448/2020\|EPI_ISL_738752\|2020- | ···················· | ···················· | ···················· | ···················· | ···················· | ···················· |
| Z | 2590 | hCoV-19/USA/CA-CZB-14124/2020\|EPI_ISL_738753\|2020- | ···················· | ···················· | ···················· | ···················· | ···················· | ···················· |
| Z | 2591 | hCoV-19/USA/CA-CZB-14602/2020\|EPI_ISL_738754\|2020- | ···················· | ···················· | ···················· | ···················· | ···················· | ···················· |
| Z | 2592 | hCoV-19/USA/CA-CZB-14666/2020\|EPI_ISL_738755\|2020- | ···················· | ···················· | ···················· | ···················· | ···················· | ···················· |
| Z | 2593 | hCoV-19/USA/CA-CZB-14749/2020\|EPI_ISL_738756\|2020- | ···················· | ···················· | ···················· | ···················· | ···················· | ···················· |
| Z | 2594 | hCoV-19/USA/CA-CZB-14210/2020\|EPI_ISL_738757\|2020- | ···················· | ·············A······ | ···················· | ···················· | ···················· | ···················· |
| Z | 2595 | hCoV-19/USA/CA-CZB-15425/2020\|EPI_ISL_738758\|2020- | ···················· | ·A·················· | ···················· | ···················· | ···················· | ···················· |
| Z | 2596 | hCoV-19/USA/CA-CZB-14781/2020\|EPI_ISL_738759\|2020- | ···················· | ···················· | ···················· | ···················· | ···················· | ···················· |
| Z | 2597 | hCoV-19/USA/CA-CZB-13990/2020\|EPI_ISL_738760\|2020- | ···················· | ···················· | ···················· | ···················· | ···················· | ···················· |
| Z | 2598 | hCoV-19/USA/CA-CZB-14644/2020\|EPI_ISL_738761\|2020- | ···················· | ···················· | ···················· | ···················· | ···················· | ···················· |
| Z | 2600 | hCoV-19/USA/CA-CZB-15145/2020\|EPI_ISL_738763\|2020- | ···················· | ·A·················· | ···················· | ···················· | ···················· | ···················· |
| Z | 2601 | hCoV-19/USA/CA-CZB-15236/2020\|EPI_ISL_738764\|2020- | ···················· | ···················· | ···················· | ···················· | ···················· | ···················· |
| Z | 2606 | hCoV-19/USA/CA-CZB-15008/2020\|EPI_ISL_738769\|2020- | ···················· | ·A·················· | ···················· | ···················· | ···················· | ···················· |
| Z | 2607 | hCoV-19/USA/CA-CZB-16169/2020\|EPI_ISL_738770\|2020- | ···················· | ·A·················· | ···················· | ···················· | ···················· | ···················· |
| Z | 2608 | hCoV-19/USA/CA-CZB-14995/2020\|EPI_ISL_738771\|2020- | ···················· | ···················· | ···················· | ···················· | ···················· | ···················· |
| Z | 2609 | hCoV-19/USA/CA-CZB-15108/2020\|EPI_ISL_738772\|2020- | ···················· | ···················· | ···················· | ···················· | N··················· | ···················· |
| Z | 2610 | hCoV-19/USA/CA-CZB-14113/2020\|EPI_ISL_738773\|2020- | ···················· | ···················· | ···················· | ···················· | ···················· | ···················· |
| Z | 2613 | hCoV-19/USA/CA-CZB-13884/2020\|EPI_ISL_738776\|2020- | ···················· | ···················· | ···················· | ···················· | ···················· | ···················· |
| Z | 2614 | hCoV-19/USA/CA-CZB-11277/2020\|EPI_ISL_738777\|2020- | ···················· | ···················· | ···················· | ···················· | ···················· | ···················· |
| Z | 2615 | hCoV-19/USA/CA-CZB-14716/2020\|EPI_ISL_738778\|2020- | ···················· | ·A·················· | ···················· | ···················· | ···················· | ···················· |
| Z | 2616 | hCoV-19/USA/CA-CZB-14783/2020\|EPI_ISL_738779\|2020- | ···················· | ···················· | ···················· | ···················· | ···················· | ···················· |
| Z | 2617 | hCoV-19/USA/CA-CZB-15001/2020\|EPI_ISL_738780\|2020- | ···················· | ···················· | ···················· | ···················· | ···················· | ···················· |
| Z | 2619 | hCoV-19/USA/CA-CZB-14921/2020\|EPI_ISL_738782\|2020- | ···················· | ···················· | ···················· | ···················· | ···················· | ···················· |
| Z | 2620 | hCoV-19/USA/CA-CZB-13833/2020\|EPI_ISL_738783\|2020- | ···················· | ···················· | ···················· | ···················· | ···················· | ···················· |
| Z | 2621 | hCoV-19/USA/CA-CZB-15424/2020\|EPI_ISL_738784\|2020- | ···················· | ···················· | ···················· | ···················· | ···················· | ···················· |
| Z | 2622 | hCoV-19/USA/CA-CZB-14715/2020\|EPI_ISL_738785\|2020- | ···················· | ·A·················· | ···················· | ···················· | ···················· | ···················· |
| Z | 2623 | hCoV-19/USA/CA-CZB-14766/2020\|EPI_ISL_738786\|2020- | ···················· | ···················· | ···················· | ···················· | ···················· | ···················· |
| Z | 2624 | hCoV-19/USA/CA-CZB-15455/2020\|EPI_ISL_738787\|2020- | ···················· | ···················· | ···················· | ···················· | ···················· | ···················· |
| Z | 2625 | hCoV-19/USA/CA-CZB-14134/2020\|EPI_ISL_738788\|2020- | ···················· | ···················· | ···················· | ···················· | ···················· | ···················· |
| Z | 2626 | hCoV-19/USA/CA-CZB-15175/2020\|EPI_ISL_738789\|2020- | ···················· | ·A·················· | ···················· | ···················· | ···················· | ···················· |
| Z | 2627 | hCoV-19/USA/CA-CZB-14879/2020\|EPI_ISL_738790\|2020- | ···················· | ···················· | ···················· | ···················· | ···················· | ···················· |
| Z | 2628 | hCoV-19/USA/CA-CZB-15073/2020\|EPI_ISL_738791\|2020- | ···················· | ···················· | ···················· | ···················· | ···················· | ···················· |
| Z | 2632 | hCoV-19/USA/CA-CZB-14779/2020\|EPI_ISL_738795\|2020- | ···················· | ···················· | ···················· | ···················· | ···················· | ···················· |
| Z | 2634 | hCoV-19/USA/CA-CZB-14139/2020\|EPI_ISL_738797\|2020- | ···················· | ·A·················· | ···················· | ···················· | ···················· | ···················· |
| Z | 2636 | hCoV-19/USA/CA-CZB-14639/2020\|EPI_ISL_738799\|2020- | ···················· | ···················· | ···················· | ···················· | ···················· | ···················· |
| Z | 2638 | hCoV-19/USA/CA-CZB-14864/2020\|EPI_ISL_739235\|2020- | ···················· | ···················· | ···················· | ···················· | ···················· | ···················· |
| Z | 2640 | hCoV-19/USA/CA-CZB-15075/2020\|EPI_ISL_739237\|2020- | ···················· | ···················· | ···················· | ···················· | ···················· | ···················· |
| Z | 2641 | hCoV-19/USA/CA-CZB-14636/2020\|EPI_ISL_739238\|2020- | ···················· | ···················· | ···················· | ···················· | ···················· | ···················· |
| Z | 2642 | hCoV-19/USA/CA-CZB-14213/2020\|EPI_ISL_739239\|2020- | ···················· | ·············A······ | ···················· | ···················· | ···················· | ···················· |
| Z | 2643 | hCoV-19/Luxembourg/LNS2902167/2020\|EPI_ISL_740214\| | ···················· | ···················· | ···················· | ···················· | ···················· | ···················· |
| Z | 2644 | hCoV-19/Luxembourg/LNS2904387/2020\|EPI_ISL_740215\| | ···················· | ···················· | ···················· | ···················· | ···················· | ···················· |
| Z | 2645 | hCoV-19/Luxembourg/LNS2905692/2020\|EPI_ISL_740216\| | ···················· | ···················· | ···················· | ···················· | ···················· | ···················· |
| Z | 2646 | hCoV-19/Luxembourg/LNS2914409/2020\|EPI_ISL_740217\| | ···················· | ···················· | ···················· | ···················· | ···················· | ···················· |
| Z | 2647 | hCoV-19/Luxembourg/LNS2937123/2020\|EPI_ISL_740220\| | ···················· | ···················· | ···················· | ···················· | ···················· | ···················· |
| Z | 2648 | hCoV-19/Luxembourg/LNS2946754/2020\|EPI_ISL_740221\| | ···················· | ···················· | ···················· | ···················· | ···················· | ···················· |
| Z | 2649 | hCoV-19/Luxembourg/LNS2951750/2020\|EPI_ISL_740222\| | ···················· | ···················· | ···················· | ···················· | ···················· | ···················· |
| Z | 2650 | hCoV-19/Luxembourg/LNS2955439/2020\|EPI_ISL_740223\| | ···················· | ···················· | ···················· | ···················· | ···················· | ···················· |
| Z | 2651 | hCoV-19/Luxembourg/LNS2958675/2020\|EPI_ISL_740224\| | ···················· | ···················· | ···················· | ···················· | ···················· | ···················· |
| Z | 2652 | hCoV-19/England/NORW-222EDF/2020\|EPI_ISL_741545\|20 | ···················· | ···················· | ···················· | ···················· | ···················· | ···················· |
| Z | 2653 | hCoV-19/England/NORW-222EEE/2020\|EPI_ISL_741546\|20 | ···················· | ···················· | ···················· | ···················· | ···················· | ···················· |
| Z | 2654 | hCoV-19/England/NORW-222EFD/2020\|EPI_ISL_741547\|20 | ···················· | ·A·················· | ···················· | ···················· | ···················· | ··A················· |
| Z | 2655 | hCoV-19/England/NORW-222F09/2020\|EPI_ISL_741548\|20 | ···················· | ···················· | ···················· | ···················· | ···················· | ···················· |
| Z | 2656 | hCoV-19/England/NORW-222F27/2020\|EPI_ISL_741550\|20 | ···················· | ···················· | ···················· | ···················· | ···················· | ···················· |
| Z | 2657 | hCoV-19/England/NORW-222F36/2020\|EPI_ISL_741551\|20 | ···················· | ···················· | ···················· | ···················· | ···················· | ···················· |
| Z | 2658 | hCoV-19/England/NORW-222F45/2020\|EPI_ISL_741552\|20 | ···················· | ···················· | ···················· | ···················· | ···················· | ···················· |
| Z | 2659 | hCoV-19/England/NORW-222F54/2020\|EPI_ISL_741553\|20 | ···················· | ···················· | ···················· | ···················· | ···················· | ···················· |
| Z | 2660 | hCoV-19/England/NORW-222F72/2020\|EPI_ISL_741554\|20 | ···················· | ···················· | ···················· | ···················· | ···················· | ···················· |
| Z | 2661 | hCoV-19/England/NORW-222F90/2020\|EPI_ISL_741555\|20 | ···················· | ···················· | ···················· | ···················· | ···················· | ···················· |
| Z | 2663 | hCoV-19/Wales/PHWC-49DB21/2020\|EPI_ISL_742877\|2020 | ···················· | ···················· | ···················· | ···················· | ···················· | ···················· |
| Z | 2664 | hCoV-19/Wales/PHWC-49DB4F/2020\|EPI_ISL_742879\|2020 | ···················· | ···················· | ···················· | ···················· | ···················· | ···················· |

| | | | | | | | | |
|---|---|---|---|---|---|---|---|---|
| Z | 2665 | hCoV-19/Wales/PHWC-49DBB8/2020\|EPI_ISL_742880\|2020 | ···················· | ···················· | ···················· | ···················· | ···················· | ···················· |
| Z | 2666 | hCoV-19/Wales/PHWC-49DBC7/2020\|EPI_ISL_742881\|2020 | ···················· | ···················· | ···················· | ···················· | ···················· | ···················· |
| Z | 2667 | hCoV-19/Wales/PHWC-49DBE5/2020\|EPI_ISL_742883\|2020 | ···················· | ···················· | ···················· | ···················· | ···················· | ···················· |
| Z | 2670 | hCoV-19/Wales/PHWC-49DC1F/2020\|EPI_ISL_742886\|2020 | ···················· | ···················· | ···················· | ···················· | ···················· | ···················· |
| Z | 2671 | hCoV-19/Wales/PHWC-49F451/2020\|EPI_ISL_743217\|2020 | ···················· | ···················· | ···················· | ···················· | ···················· | ···················· |
| Z | 2672 | hCoV-19/Wales/PHWC-49F460/2020\|EPI_ISL_743218\|2020 | ···················· | ···················· | ···················· | ···················· | ···················· | ···················· |
| Z | 2673 | hCoV-19/Wales/PHWC-49F47F/2020\|EPI_ISL_743219\|2020 | ···················· | ···················· | ···················· | ···················· | ···················· | ···················· |
| Z | 2674 | hCoV-19/Wales/PHWC-49F48E/2020\|EPI_ISL_743220\|2020 | ···················· | ···················· | ···················· | ···················· | ···················· | ···················· |
| Z | 2675 | hCoV-19/Wales/PHWC-49F49D/2020\|EPI_ISL_743221\|2020 | ···················· | ···················· | ···················· | ···················· | ···················· | ···················· |
| Z | 2676 | hCoV-19/Wales/PHWC-49F4AC/2020\|EPI_ISL_743222\|2020 | ···················· | ···················· | ···················· | ···················· | ···················· | ···················· |
| Z | 2678 | hCoV-19/Wales/PHWC-49F4CA/2020\|EPI_ISL_743224\|2020 | ···················· | ···················· | ···················· | ···················· | ···················· | ···················· |
| Z | 2679 | hCoV-19/Wales/PHWC-49F4E8/2020\|EPI_ISL_743226\|2020 | ···················· | ···················· | ···················· | ···················· | ···················· | ···················· |
| Z | 2680 | hCoV-19/Wales/PHWC-49F4F7/2020\|EPI_ISL_743227\|2020 | ···················· | ···················· | ···················· | ···················· | ···················· | ···················· |
| Z | 2681 | hCoV-19/Wales/PHWC-49F503/2020\|EPI_ISL_743228\|2020 | ···················· | ···················· | ···················· | ···················· | ···················· | ···················· |
| Z | 2682 | hCoV-19/Wales/PHWC-49F512/2020\|EPI_ISL_743229\|2020 | ···················· | ···················· | ···················· | ···················· | ···················· | ···················· |
| Z | 2683 | hCoV-19/Wales/PHWC-49F521/2020\|EPI_ISL_743230\|2020 | ···················· | ···················· | ···················· | ···················· | ···················· | ···················· |
| Z | 2684 | hCoV-19/Wales/PHWC-49F530/2020\|EPI_ISL_743231\|2020 | ···················· | ···················· | ···················· | ···················· | ···················· | ···················· |
| Z | 2685 | hCoV-19/Wales/PHWC-49F54F/2020\|EPI_ISL_743232\|2020 | ···················· | ···················· | ···················· | ···················· | ···················· | ···················· |
| Z | 2687 | hCoV-19/Wales/PHWC-49F57C/2020\|EPI_ISL_743234\|2020 | ···················· | ···················· | ···················· | ···················· | ···················· | ···················· |
| Z | 2689 | hCoV-19/Wales/PHWC-49F5B8/2020\|EPI_ISL_743237\|2020 | ···················· | ···················· | ···················· | ···················· | ···················· | ···················· |
| Z | 2690 | hCoV-19/Wales/PHWC-49F5C7/2020\|EPI_ISL_743238\|2020 | ···················· | ······A············· | ···················· | ···················· | ···················· | ···················· |
| Z | 2691 | hCoV-19/Wales/PHWC-49F5D6/2020\|EPI_ISL_743239\|2020 | ···················· | ···················· | ···················· | ···················· | ···················· | ···················· |
| Z | 2693 | hCoV-19/Wales/PHWC-49F5F4/2020\|EPI_ISL_743241\|2020 | ···················· | ···················· | ···················· | ···················· | ···················· | ···················· |
| Z | 2694 | hCoV-19/Wales/PHWC-49F63D/2020\|EPI_ISL_743245\|2020 | ···················· | ···················· | ···················· | ···················· | ···················· | ···················· |
| Z | 2695 | hCoV-19/Wales/PHWC-49F64C/2020\|EPI_ISL_743246\|2020 | ···················· | ···················· | ···················· | ···················· | ···················· | ···················· |
| Z | 2697 | hCoV-19/Wales/PHWC-49F679/2020\|EPI_ISL_743248\|2020 | ···················· | ···················· | ···················· | ···················· | ···················· | ···················· |
| Z | 2698 | hCoV-19/Wales/PHWC-49F688/2020\|EPI_ISL_743249\|2020 | ···················· | ···················· | ···················· | ···················· | ···················· | ···················· |
| Z | 2699 | hCoV-19/Wales/PHWC-49F6A6/2020\|EPI_ISL_743251\|2020 | ···················· | ···················· | ···················· | ···················· | ···················· | ···················· |
| Z | 2700 | hCoV-19/Wales/PHWC-49F6B5/2020\|EPI_ISL_743252\|2020 | ···················· | ···················· | ···················· | ···················· | ···················· | ···················· |
| Z | 2701 | hCoV-19/Wales/PHWC-49F6D3/2020\|EPI_ISL_743254\|2020 | ···················· | ···················· | ···················· | ···················· | ···················· | ···················· |
| Z | 2702 | hCoV-19/Wales/PHWC-49F6E2/2020\|EPI_ISL_743255\|2020 | ···················· | ···················· | ···················· | ···················· | ···················· | ···················· |
| Z | 2703 | hCoV-19/Wales/PHWC-49F70D/2020\|EPI_ISL_743257\|2020 | ···················· | ···················· | ···················· | ···················· | ···················· | ···················· |
| Z | 2704 | hCoV-19/Wales/PHWC-49F71C/2020\|EPI_ISL_743258\|2020 | ···················· | ···················· | ···················· | ···················· | ···················· | ···················· |
| Z | 2705 | hCoV-19/Wales/PHWC-49F72B/2020\|EPI_ISL_743259\|2020 | ···················· | ···················· | ···················· | ···················· | ···················· | ···················· |
| Z | 2706 | hCoV-19/Wales/PHWC-49F73A/2020\|EPI_ISL_743260\|2020 | ···················· | ···················· | ···················· | ···················· | ···················· | ···················· |
| Z | 2707 | hCoV-19/Wales/PHWC-49F749/2020\|EPI_ISL_743261\|2020 | ···················· | ···················· | ···················· | ···················· | ···················· | ···················· |
| Z | 2708 | hCoV-19/Wales/PHWC-49F758/2020\|EPI_ISL_743262\|2020 | ···················· | ···················· | ···················· | ···················· | ···················· | ···················· |
| Z | 2709 | hCoV-19/Wales/PHWC-49F767/2020\|EPI_ISL_743263\|2020 | ···················· | ···················· | ···················· | ···················· | ···················· | ···················· |
| Z | 2710 | hCoV-19/Wales/PHWC-49F776/2020\|EPI_ISL_743264\|2020 | ···················· | ···················· | ···················· | ···················· | ···················· | ···················· |
| Z | 2711 | hCoV-19/Wales/PHWC-49F785/2020\|EPI_ISL_743265\|2020 | ···················· | ···················· | ···················· | ···················· | ···················· | ···················· |
| Z | 2712 | hCoV-19/Wales/PHWC-49F794/2020\|EPI_ISL_743266\|2020 | ···················· | ···················· | ···················· | ···················· | ···················· | ···················· |
| Z | 2713 | hCoV-19/Wales/PHWC-49F7B2/2020\|EPI_ISL_743268\|2020 | ···················· | ···················· | ···················· | ···················· | ···················· | ···················· |
| Z | 2714 | hCoV-19/Wales/PHWC-49F7C1/2020\|EPI_ISL_743269\|2020 | ···················· | ···················· | ···················· | ···················· | ···················· | ···················· |
| Z | 2715 | hCoV-19/Wales/PHWC-49F7D0/2020\|EPI_ISL_743270\|2020 | ···················· | ···················· | ···················· | ···················· | ···················· | ···················· |
| Z | 2716 | hCoV-19/Wales/PHWC-49F7EF/2020\|EPI_ISL_743271\|2020 | ···················· | ···················· | ···················· | ···················· | ···················· | ···················· |
| Z | 2717 | hCoV-19/Wales/PHWC-49F7FE/2020\|EPI_ISL_743272\|2020 | ···················· | ···················· | ···················· | ···················· | ···················· | ···················· |
| Z | 2718 | hCoV-19/Wales/PHWC-49F80A/2020\|EPI_ISL_743273\|2020 | ···················· | ······A············· | ···················· | ···················· | ···················· | ···················· |
| Z | 2719 | hCoV-19/Wales/PHWC-49F819/2020\|EPI_ISL_743274\|2020 | ···················· | ···················· | ···················· | ···················· | ···················· | ···················· |
| Z | 2720 | hCoV-19/Wales/PHWC-49F828/2020\|EPI_ISL_743275\|2020 | ···················· | ···················· | ···················· | ···················· | ···················· | ···················· |
| Z | 2721 | hCoV-19/Wales/PHWC-49F837/2020\|EPI_ISL_743276\|2020 | ···················· | ···················· | ···················· | ···················· | ···················· | ···················· |
| Z | 2722 | hCoV-19/Wales/PHWC-49F855/2020\|EPI_ISL_743278\|2020 | ···················· | ···················· | ···················· | ···················· | ···················· | ···················· |
| Z | 2723 | hCoV-19/Wales/PHWC-49F864/2020\|EPI_ISL_743279\|2020 | ···················· | ···················· | ···················· | ···················· | ···················· | ···················· |
| Z | 2724 | hCoV-19/Wales/PHWC-49F882/2020\|EPI_ISL_743281\|2020 | ···················· | ···················· | ···················· | ···················· | ···················· | ···················· |
| Z | 2725 | hCoV-19/Wales/PHWC-49F891/2020\|EPI_ISL_743282\|2020 | ···················· | ···················· | ···················· | ···················· | ···················· | ···················· |
| Z | 2726 | hCoV-19/Wales/PHWC-49F8A0/2020\|EPI_ISL_743283\|2020 | ···················· | ···················· | ···················· | ···················· | ···················· | ···················· |
| Z | 2727 | hCoV-19/Wales/PHWC-49F8BF/2020\|EPI_ISL_743284\|2020 | ···················· | ···················· | ···················· | ···················· | ···················· | ···A················ |
| Z | 2728 | hCoV-19/Wales/PHWC-49F8CE/2020\|EPI_ISL_743285\|2020 | ···················· | ···················· | ···················· | ···················· | ···················· | ···················· |
| Z | 2729 | hCoV-19/Wales/PHWC-49F8DD/2020\|EPI_ISL_743286\|2020 | ···················· | ···················· | ···················· | ···················· | ···················· | ···················· |
| Z | 2730 | hCoV-19/Wales/PHWC-49F8EC/2020\|EPI_ISL_743287\|2020 | ···················· | ···················· | ···················· | ···················· | ···················· | ···················· |
| Z | 2731 | hCoV-19/Wales/PHWC-49F907/2020\|EPI_ISL_743289\|2020 | ···················· | ···················· | ···················· | ···················· | ···················· | ···················· |
| Z | 2732 | hCoV-19/Wales/PHWC-49F916/2020\|EPI_ISL_743290\|2020 | ···················· | ···················· | ···················· | ···················· | ···················· | ···················· |
| Z | 2733 | hCoV-19/Wales/PHWC-49F925/2020\|EPI_ISL_743291\|2020 | ···················· | ···················· | ···················· | ···················· | ···················· | ···················· |
| Z | 2734 | hCoV-19/Wales/PHWC-49F934/2020\|EPI_ISL_743292\|2020 | ···················· | ···················· | ···················· | ···················· | ···················· | ···················· |
| Z | 2735 | hCoV-19/Wales/PHWC-49F952/2020\|EPI_ISL_743294\|2020 | ···················· | ···················· | ···················· | ···················· | ···················· | ···················· |
| Z | 2736 | hCoV-19/Wales/PHWC-49F961/2020\|EPI_ISL_743295\|2020 | ···················· | ···················· | ···················· | ···················· | ···················· | ···················· |
| Z | 2737 | hCoV-19/Wales/PHWC-49F970/2020\|EPI_ISL_743296\|2020 | ···················· | ···················· | ···················· | ···················· | ···················· | ···················· |
| Z | 2738 | hCoV-19/Wales/PHWC-49F98F/2020\|EPI_ISL_743297\|2020 | ···················· | ···················· | ···················· | ···················· | ···················· | ···················· |
| Z | 2739 | hCoV-19/Wales/PHWC-49F99E/2020\|EPI_ISL_743298\|2020 | ···················· | ···················· | ···················· | ···················· | ···················· | ···················· |
| Z | 2740 | hCoV-19/Wales/PHWC-49F9AD/2020\|EPI_ISL_743299\|2020 | ·T·················· | ···················· | ···················· | ···················· | ···················· | ···················· |
| Z | 2741 | hCoV-19/Luxembourg/LNS7064269/2020\|EPI_ISL_744504\| | ···················· | ···················· | ···················· | ···················· | ···················· | ···················· |
| Z | 2742 | hCoV-19/Luxembourg/LNS7066582/2020\|EPI_ISL_744505\| | ···················· | ···················· | ···················· | ···················· | ···················· | ···················· |
| Z | 2743 | hCoV-19/Luxembourg/LNS7078792/2020\|EPI_ISL_744506\| | ···················· | ···················· | ···················· | ···················· | ···················· | ···················· |
| Z | 2744 | hCoV-19/Luxembourg/LNS7081939/2020\|EPI_ISL_744507\| | ···················· | ···················· | ···················· | ···················· | ···················· | ···················· |
| Z | 2745 | hCoV-19/Luxembourg/LNS7083545/2020\|EPI_ISL_744508\| | ···················· | ···················· | ···················· | ···················· | ···················· | ···················· |
| Z | 2746 | hCoV-19/Luxembourg/LNS7083587/2020\|EPI_ISL_744509\| | ···················· | ···················· | ···················· | ···················· | ···················· | ···················· |
| Z | 2747 | hCoV-19/Luxembourg/LNS7305473/2020\|EPI_ISL_744548\| | ···················· | ···················· | ···················· | ···················· | ···················· | ···················· |
| Z | 2748 | hCoV-19/Luxembourg/LNS7305819/2020\|EPI_ISL_744549\| | ···················· | ···················· | ···················· | ···················· | ···················· | ···················· |
| Z | 2749 | hCoV-19/Luxembourg/LNS7312611/2020\|EPI_ISL_744550\| | ···················· | ···················· | ···················· | ···················· | ···················· | ···················· |
| Z | 2750 | hCoV-19/Luxembourg/LNS7322663/2020\|EPI_ISL_744552\| | ···················· | ···················· | ···················· | ···················· | ···················· | ···················· |
| Z | 2751 | hCoV-19/Luxembourg/LNS7330392/2020\|EPI_ISL_744553\| | ···················· | ···················· | ···················· | ···················· | ···················· | ···················· |

| | | | | | | | | |
|---|---|---|---|---|---|---|---|---|
| Z | 2752 hCoV-19/Luxembourg/LNS7333005/2020\|EPI_ISL_744554\| | ···················· | ···················· | ····················· | ···················· | ······T············· | ···················· | |
| Z | 2753 hCoV-19/Luxembourg/LNS7339710/2020\|EPI_ISL_744555\| | ···················· | ···················· | ····················· | ···················· | ···················· | ···················· | |
| Z | 2754 hCoV-19/Luxembourg/LNS7350759/2020\|EPI_ISL_744556\| | ···················· | ···················· | ····················· | ···················· | ···················· | ···················· | |
| Z | 2755 hCoV-19/Luxembourg/LNS7357799/2020\|EPI_ISL_744557\| | ···················· | ···················· | ····················· | ···················· | ···················· | ···················· | |
| Z | 2756 hCoV-19/Luxembourg/LNS7385509/2020\|EPI_ISL_744558\| | ···················· | ···················· | ····················· | ···················· | ···················· | ···················· | |
| Z | 2757 hCoV-19/Luxembourg/LNS7385935/2020\|EPI_ISL_744559\| | ···················· | ···················· | ····················· | ···················· | ···················· | ···················· | |
| Z | 2758 hCoV-19/Luxembourg/LNS7392121/2020\|EPI_ISL_744563\| | ···················· | ···················· | ····················· | ···················· | ···················· | ···················· | |
| Z | 2759 hCoV-19/Luxembourg/LNS7394413/2020\|EPI_ISL_744564\| | ···················· | ···················· | ····················· | ···················· | ···················· | ···················· | |
| Z | 2760 hCoV-19/Luxembourg/LNS7395205/2020\|EPI_ISL_744565\| | ···················· | ···················· | ····················· | ···················· | ···················· | ···················· | |
| Z | 2761 hCoV-19/Luxembourg/LNS7397711/2020\|EPI_ISL_744566\| | ···········T········ | ···················· | ····················· | ···················· | ···················· | ···················· | |
| Z | 2762 hCoV-19/Luxembourg/LNS7409231/2020\|EPI_ISL_744567\| | ···················· | ···················· | ····················· | ···················· | ···················· | ···················· | |
| Z | 2763 hCoV-19/Luxembourg/LNS7418259/2020\|EPI_ISL_744568\| | ···················· | ···················· | ····················· | ···················· | ···················· | ···················· | |
| Z | 2764 hCoV-19/Luxembourg/LNS7420446/2020\|EPI_ISL_744569\| | ···················· | ···················· | ····················· | ···················· | ···················· | ···················· | |
| Z | 2765 hCoV-19/Luxembourg/LNS7427655/2020\|EPI_ISL_744570\| | ···················· | ···················· | ····················· | ···················· | ···················· | ···················· | |
| Z | 2766 hCoV-19/Luxembourg/LNS7437554/2020\|EPI_ISL_744571\| | ···················· | ···················· | ····················· | ···················· | ···················· | ···················· | |
| Z | 2767 hCoV-19/Luxembourg/LNS7459401/2020\|EPI_ISL_744572\| | ···················· | ···················· | ····················· | ···················· | ···················· | ···················· | |
| Z | 2768 hCoV-19/Luxembourg/LNS7460064/2020\|EPI_ISL_744573\| | ···················· | ···················· | ····················· | ···················· | ···················· | ···················· | |
| Z | 2769 hCoV-19/Luxembourg/LNS7465046/2020\|EPI_ISL_744574\| | ···················· | ···················· | ····················· | ···················· | ···················· | ···················· | |
| Z | 2770 hCoV-19/Luxembourg/LNS7469233/2020\|EPI_ISL_744575\| | ···················· | ···················· | ····················· | ···················· | ······T············· | ···················· | |
| Z | 2771 hCoV-19/Luxembourg/LNS7492747/2020\|EPI_ISL_744576\| | ···················· | ···················· | ····················· | ···················· | ···················· | ···················· | |
| Z | 2772 hCoV-19/Luxembourg/LNS7493897/2020\|EPI_ISL_744577\| | ···················· | ···················· | ····················· | ···················· | ···················· | ···················· | |
| Z | 2773 hCoV-19/Luxembourg/LNS7495788/2020\|EPI_ISL_744578\| | ···················· | ···················· | ····················· | ···················· | ···················· | ···················· | |
| Z | 2774 hCoV-19/Luxembourg/LNS7496221/2020\|EPI_ISL_744579\| | ···················· | ···················· | ····················· | ···················· | ···················· | ···················· | |
| Z | 2775 hCoV-19/Luxembourg/LNS7510950/2020\|EPI_ISL_744580\| | ···················· | G··················· | ····················· | ···················· | ···················· | ···················· | |
| Z | 2776 hCoV-19/Luxembourg/LNS7512526/2020\|EPI_ISL_744581\| | ···················· | ···················· | ····················· | ···················· | ···················· | ···················· | |
| Z | 2777 hCoV-19/Luxembourg/LNS7516199/2020\|EPI_ISL_744582\| | ···················· | ···················· | ····················· | ···················· | ···················· | ···················· | |
| Z | 2778 hCoV-19/Luxembourg/LNS7532249/2020\|EPI_ISL_744583\| | ···················· | ···················· | ····················· | ···················· | ···················· | ···················· | |
| Z | 2779 hCoV-19/Luxembourg/LNS7532467/2020\|EPI_ISL_744584\| | ···················· | ···················· | ····················· | ···················· | ···················· | ···················· | |
| Z | 2780 hCoV-19/Luxembourg/LNS7550441/2020\|EPI_ISL_744586\| | ···················· | ···················· | ····················· | ···················· | ···················· | ···················· | |
| Z | 2781 hCoV-19/Luxembourg/LNS7566407/2020\|EPI_ISL_744587\| | ···················· | ···················· | ····················· | ···················· | ···················· | ···················· | |
| Z | 2782 hCoV-19/Luxembourg/LNS7579436/2020\|EPI_ISL_744589\| | ···················· | ···················· | ····················· | ···················· | ···················· | ···················· | |
| Z | 2783 hCoV-19/Luxembourg/LNS7580348/2020\|EPI_ISL_744590\| | ···················· | ···················· | ····················· | ···················· | ···················· | ···················· | |
| Z | 2784 hCoV-19/Luxembourg/LNS7584484/2020\|EPI_ISL_744591\| | ···················· | ···················· | ····················· | ···················· | ···················· | ···················· | |
| Z | 2785 hCoV-19/Luxembourg/LNS7586283/2020\|EPI_ISL_744592\| | ···················· | ···················· | ····················· | ···················· | ···················· | ···················· | |
| Z | 2786 hCoV-19/Luxembourg/LNS7588118/2020\|EPI_ISL_744593\| | ···················· | ···················· | ····················· | ···················· | ···················· | ···················· | |
| Z | 2787 hCoV-19/Luxembourg/LNS7593111/2020\|EPI_ISL_744594\| | ···················· | ···················· | ····················· | ···················· | ···················· | ···················· | |
| Z | 2788 hCoV-19/Luxembourg/LNS7601469/2020\|EPI_ISL_744596\| | ···················· | ···················· | ····················· | ···················· | ···················· | ···················· | |
| Z | 2789 hCoV-19/Luxembourg/LNS7617595/2020\|EPI_ISL_744599\| | ···················· | ···················· | ····················· | ···················· | ···················· | ···················· | |
| Z | 2790 hCoV-19/USA/UT-UPHL-2012890972/2020\|EPI_ISL_745835 | ···················· | ···················· | ····················· | ···················· | ···················· | ···················· | |
| Z | 2791 hCoV-19/USA/UT-UPHL-2012756176/2020\|EPI_ISL_745836 | ···················· | ·A·················· | ····················· | ···················· | ···················· | ···················· | |
| Z | 2792 hCoV-19/USA/UT-UPHL-2012073870/2020\|EPI_ISL_745837 | ···················· | ·A·················· | ····················· | ···················· | ···················· | ···················· | |
| Z | 2793 hCoV-19/USA/UT-UPHL-2012117735/2020\|EPI_ISL_745838 | ···················· | ·A·················· | ····················· | ···················· | ···················· | ···················· | |
| Z | 2794 hCoV-19/USA/UT-UPHL-2012376972/2020\|EPI_ISL_745839 | ···················· | ···················· | ····················· | ···················· | ···················· | ···················· | |
| Z | 2795 hCoV-19/USA/UT-UPHL-2012742341/2020\|EPI_ISL_745843 | ···················· | ···················· | ····················· | ···················· | ···················· | ···················· | |
| Z | 2796 hCoV-19/USA/UT-UPHL-2012972043/2020\|EPI_ISL_745879 | ···················· | ···················· | ····················· | ···················· | ···················· | ···················· | |
| Z | 2797 hCoV-19/USA/UT-UPHL-2012338415/2020\|EPI_ISL_745880 | ···················· | ···················· | ····················· | ···················· | ···················· | ···················· | |
| Z | 2798 hCoV-19/USA/UT-UPHL-2012620468/2020\|EPI_ISL_745881 | ···················· | ···················· | ····················· | ···················· | ···················· | ···················· | |
| Z | 2799 hCoV-19/USA/UT-UPHL-2012492144/2020\|EPI_ISL_745882 | ···················· | ···················· | ····················· | ···················· | ···················· | ···················· | |
| Z | 2800 hCoV-19/USA/UT-UPHL-2012666296/2020\|EPI_ISL_745883 | ···················· | ·A·················· | ····················· | ···················· | ···················· | ···················· | |
| Z | 2801 hCoV-19/USA/UT-UPHL-2012414957/2020\|EPI_ISL_745884 | ···················· | ···················· | ····················· | ···················· | ···················· | ···················· | |
| Z | 2802 hCoV-19/USA/UT-UPHL-2012041204/2020\|EPI_ISL_745885 | ···················· | ···················· | ····················· | ···················· | ···················· | ···················· | |
| Z | 2803 hCoV-19/USA/UT-UPHL-2012380818/2020\|EPI_ISL_745886 | ···················· | ···················· | ····················· | ···················· | ···················· | ···················· | |
| Z | 2804 hCoV-19/USA/UT-UPHL-2012464023/2020\|EPI_ISL_745887 | ···················· | ···················· | ····················· | ···················· | ···················· | ···················· | |
| Z | 2805 hCoV-19/USA/UT-UPHL-2012085638/2020\|EPI_ISL_745888 | ···················· | ···················· | ····················· | ···················· | ···················· | ···················· | |
| Z | 2806 hCoV-19/USA/UT-UPHL-2012543390/2020\|EPI_ISL_745889 | ···················· | ·A·················· | ····················· | ···················· | ···················· | ···················· | |
| Z | 2807 hCoV-19/USA/UT-UPHL-2012442845/2020\|EPI_ISL_745890 | ···················· | ···················· | ····················· | ···················· | ···················· | ···················· | |
| Z | 2808 hCoV-19/USA/UT-UPHL-2012819923/2020\|EPI_ISL_745891 | ···················· | ·A·················· | ····················· | ···················· | ···················· | ···················· | |
| Z | 2809 hCoV-19/USA/UT-UPHL-2012337841/2020\|EPI_ISL_745892 | ···················· | ···················· | ····················· | ···················· | ···················· | ···················· | |
| Z | 2810 hCoV-19/USA/UT-UPHL-2012000183/2020\|EPI_ISL_745893 | ···················· | ·A·················· | ····················· | ···················· | ···················· | ···················· | |
| Z | 2811 hCoV-19/USA/UT-UPHL-2012780736/2020\|EPI_ISL_745894 | ···················· | ···················· | ····················· | ···················· | ···················· | ···················· | |
| Z | 2812 hCoV-19/USA/UT-UPHL-2012896384/2020\|EPI_ISL_745895 | ···················· | ···················· | ····················· | ···················· | ···················· | ···················· | |
| Z | 2813 hCoV-19/USA/UT-UPHL-2012639635/2020\|EPI_ISL_745896 | ···················· | ·A·················· | ····················· | ···················· | ···················· | ···················· | |
| Z | 2814 hCoV-19/USA/UT-UPHL-2012768135/2020\|EPI_ISL_745897 | ···················· | ···················· | ····················· | ···················· | ···················· | ···················· | |
| Z | 2815 hCoV-19/USA/UT-UPHL-2012124268/2020\|EPI_ISL_745898 | ···················· | ·A·················· | ····················· | ···················· | ···················· | ···················· | |
| Z | 2816 hCoV-19/USA/UT-UPHL-2012218419/2020\|EPI_ISL_745899 | ···················· | ···················· | ····················· | ···················· | ···················· | ···················· | |
| Z | 2818 hCoV-19/USA/CA-CZB-13959/2020\|EPI_ISL_739201\|2020- | ···················· | ···················· | ····················· | ···················· | ···················· | ···················· | |
| Z | 2819 hCoV-19/USA/CA-CZB-13779/2020\|EPI_ISL_739202\|2020- | ···················· | ·A·················· | ····················· | ···················· | NNN················· | ···················· | 1 |
| Z | 2820 hCoV-19/USA/CA-CZB-14590/2020\|EPI_ISL_739203\|2020- | ···················· | ···················· | ····················· | ···················· | ···················· | ···················· | |
| Z | 2821 hCoV-19/USA/CA-CZB-14575/2020\|EPI_ISL_739204\|2020- | ···················· | ·A·················· | ····················· | ···················· | ···················· | ···················· | |
| Z | 2822 hCoV-19/USA/CA-CZB-14684/2020\|EPI_ISL_739205\|2020- | ···················· | ···················· | ····················· | ···················· | ···················· | ···················· | |
| Z | 2823 hCoV-19/USA/CA-CZB-13942/2020\|EPI_ISL_739206\|2020- | ···················· | ···················· | ····················· | ···················· | ···················· | ···················· | |
| Z | 2825 hCoV-19/USA/CA-CZB-13975/2020\|EPI_ISL_739208\|2020- | ···················· | ···················· | ····················· | ···················· | ···················· | ···················· | |
| Z | 2826 hCoV-19/USA/CA-CZB-14421/2020\|EPI_ISL_739209\|2020- | ···················· | ···················· | ····················· | ···················· | ···················· | ···················· | |
| Z | 2827 hCoV-19/USA/CA-CZB-14799/2020\|EPI_ISL_739210\|2020- | ···················· | ···················· | ····················· | ···················· | ···················· | ···················· | |
| Z | 2828 hCoV-19/USA/CA-CZB-14882/2020\|EPI_ISL_739211\|2020- | ···················· | ···················· | ····················· | ···················· | ···················· | ···················· | |
| Z | 2830 hCoV-19/USA/CA-CZB-13866/2020\|EPI_ISL_739213\|2020- | ···················· | ···················· | ····················· | ···················· | ···················· | ···················· | |
| Z | 2831 hCoV-19/USA/CA-CZB-15285/2020\|EPI_ISL_739214\|2020- | ···················· | ···················· | ····················· | ···················· | ···················· | ···················· | |
| Z | 2833 hCoV-19/USA/CA-CZB-14920/2020\|EPI_ISL_739216\|2020- | ···················· | ···················· | ····················· | ···················· | ···················· | ···················· | |
| Z | 2834 hCoV-19/USA/CA-CZB-14068/2020\|EPI_ISL_739217\|2020- | ···················· | ···················· | ····················· | ···················· | ···················· | ···················· | |
| Z | 2835 hCoV-19/USA/CA-CZB-14259/2020\|EPI_ISL_739218\|2020- | ···················· | ···················· | ····················· | ···················· | ···················· | ···················· | |

| | | | | | | | | |
|---|---|---|---|---|---|---|---|---|
| Z | 2837 | hCoV-19/USA/CA-CZB-13936/2020\|EPI_ISL_739220\|2020- | ···················· | ·A·················· | ····················· | ···················· | ···················· | ···················· |
| Z | 2838 | hCoV-19/USA/CA-CZB-15229/2020\|EPI_ISL_739221\|2020- | ···················· | ···················· | ····················· | ···················· | ···················· | ···················· |
| Z | 2839 | hCoV-19/USA/CA-CZB-14898/2020\|EPI_ISL_739222\|2020- | ···················· | ···················· | ····················· | ···················· | ···················· | ···················· |
| Z | 2840 | hCoV-19/USA/CA-CZB-11274/2020\|EPI_ISL_739223\|2020- | ···················· | ···················· | ····················· | ···················· | ···················· | ···················· |
| Z | 2841 | hCoV-19/USA/CA-CZB-13679/2020\|EPI_ISL_739224\|2020- | ···················· | ···················· | ····················· | ···················· | ···················· | ···················· |
| Z | 2842 | hCoV-19/USA/CA-CZB-14750/2020\|EPI_ISL_739225\|2020- | ···················· | ·A·················· | ···········T········· | ···················· | ···················· | ···················· |
| Z | 2843 | hCoV-19/USA/CA-CZB-13917/2020\|EPI_ISL_739226\|2020- | ···················· | ·A·················· | ····················· | ···················· | ···················· | ···················· |
| Z | 2845 | hCoV-19/USA/CA-CZB-14813/2020\|EPI_ISL_739228\|2020- | ···················· | ···················· | ····················· | ···················· | ···················· | ···················· |
| Z | 2846 | hCoV-19/USA/CA-CZB-14122/2020\|EPI_ISL_739229\|2020- | ···················· | ···················· | ····················· | ···················· | ···················· | ···················· |
| Z | 2847 | hCoV-19/USA/CA-CZB-14688/2020\|EPI_ISL_739230\|2020- | ···················· | ···················· | ····················· | ···················· | ···················· | ···················· |
| Z | 2848 | hCoV-19/USA/CA-CZB-14314/2020\|EPI_ISL_739231\|2020- | ···················· | ···················· | ····················· | ···················· | ···················· | ···················· |
| Z | 2850 | hCoV-19/USA/CA-CZB-14416/2020\|EPI_ISL_739233\|2020- | ···················· | ···················· | ····················· | ···················· | ···················· | ···················· |
| Z | 2851 | hCoV-19/USA/CA-CZB-14810/2020\|EPI_ISL_739240\|2020- | ···················· | ···················· | ····················· | ···················· | ···················· | ···················· |
| Z | 2852 | hCoV-19/USA/CA-CZB-14417/2020\|EPI_ISL_739241\|2020- | ···················· | ·A·················· | ····················· | ···················· | ···················· | ···················· |
| Z | 2853 | hCoV-19/USA/CA-CZB-14820/2020\|EPI_ISL_739242\|2020- | ···················· | ···················· | ····················· | ···················· | ···················· | ···················· |
| Z | 2854 | hCoV-19/USA/CA-CZB-14022/2020\|EPI_ISL_739243\|2020- | ···················· | ···················· | ····················· | ···················· | ···················· | ···················· |
| Z | 2855 | hCoV-19/USA/CA-CZB-13869/2020\|EPI_ISL_739244\|2020- | ···················· | ···················· | ····················· | ···················· | ···················· | ···················· |
| Z | 2856 | hCoV-19/USA/CA-CZB-14205/2020\|EPI_ISL_739245\|2020- | ···················· | ···················· | ····················· | ···················· | ···················· | ···················· |
| Z | 2860 | hCoV-19/USA/CA-CZB-15347/2020\|EPI_ISL_739249\|2020- | ···················· | ···················· | ····················· | ···················· | ···················· | ···················· |
| Z | 2861 | hCoV-19/USA/CA-CZB-14859/2020\|EPI_ISL_739250\|2020- | ···················· | ···················· | ····················· | ···················· | ···················· | ···················· |
| Z | 2862 | hCoV-19/USA/CA-CZB-11253/2020\|EPI_ISL_739251\|2020- | ···················· | ···················· | ····················· | ···················· | ···················· | ···················· |
| Z | 2863 | hCoV-19/USA/CA-CZB-15137/2020\|EPI_ISL_739252\|2020- | ···················· | ·A·················· | ····················· | ···················· | ···················· | ···················· |
| Z | 2864 | hCoV-19/USA/CA-CZB-15021/2020\|EPI_ISL_739253\|2020- | ···················· | ·A·················· | ····················· | ···················· | ···················· | ···················· |
| Z | 2865 | hCoV-19/USA/CA-CZB-15118/2020\|EPI_ISL_739254\|2020- | ···················· | ···················· | ····················· | ···················· | ···················· | ···················· |
| Z | 2868 | hCoV-19/USA/CA-CZB-14078/2020\|EPI_ISL_739257\|2020- | ···················· | ···················· | ····················· | ···················· | ···················· | ···················· |
| Z | 2869 | hCoV-19/USA/CA-CZB-15461/2020\|EPI_ISL_739258\|2020- | ···················· | ···················· | ···T················· | ···················· | ···················· | ···················· |
| Z | 2870 | hCoV-19/USA/CA-CZB-15435/2020\|EPI_ISL_739259\|2020- | ···················· | ···················· | ····················· | ···················· | ···················· | ···················· |
| Z | 2872 | hCoV-19/USA/CA-CZB-13901/2020\|EPI_ISL_739261\|2020- | ···················· | ·A·················· | ····················· | ···················· | ···················· | ···················· |
| Z | 2873 | hCoV-19/USA/CA-CZB-13927/2020\|EPI_ISL_739262\|2020- | ···················· | ···················· | ····················· | ···················· | ···················· | ···················· |
| Z | 2874 | hCoV-19/USA/CA-CZB-15963/2020\|EPI_ISL_739263\|2020- | ···················· | ·A·················· | ····················· | ···················· | ···················· | ···················· |
| Z | 2875 | hCoV-19/USA/CA-CZB-14722/2020\|EPI_ISL_739264\|2020- | ···················· | ···················· | ····················· | ···················· | ···················· | ···················· |
| Z | 2876 | hCoV-19/USA/CA-CZB-15472/2020\|EPI_ISL_739265\|2020- | ···················· | ···················· | ···T················· | ···················· | ···················· | ···················· |
| Z | 2877 | hCoV-19/USA/CA-CZB-14919/2020\|EPI_ISL_739266\|2020- | ···················· | ···················· | ····················· | ···················· | ···················· | ···················· |
| Z | 2879 | hCoV-19/USA/CA-CZB-14137/2020\|EPI_ISL_739268\|2020- | ···················· | ···················· | ····················· | ···················· | ···················· | ···················· |
| Z | 2880 | hCoV-19/USA/CA-CZB-14866/2020\|EPI_ISL_739269\|2020- | ···················· | ···················· | ····················· | ···················· | ···················· | ···················· |
| Z | 2881 | hCoV-19/USA/CA-CZB-14055/2020\|EPI_ISL_739270\|2020- | ···················· | ·A·················· | ····················· | ···················· | ···················· | ···················· |
| Z | 2882 | hCoV-19/USA/CA-CZB-14708/2020\|EPI_ISL_739271\|2020- | ···················· | ···················· | ····················· | ···················· | ···················· | ···················· |
| Z | 2883 | hCoV-19/USA/CA-CZB-14768/2020\|EPI_ISL_739272\|2020- | ···················· | ·A·················· | ····················· | ···················· | ···················· | ···················· |
| Z | 2884 | hCoV-19/USA/CA-CZB-15295/2020\|EPI_ISL_739273\|2020- | ···················· | ·A·················· | ····················· | ···················· | ···················· | ···················· |
| Z | 2885 | hCoV-19/USA/CA-CZB-14834/2020\|EPI_ISL_739274\|2020- | ···················· | ···················· | ····················· | ···················· | ···················· | ···················· |
| Z | 2886 | hCoV-19/USA/CA-CZB-14586/2020\|EPI_ISL_739275\|2020- | ···················· | ···················· | ····················· | ···················· | ···················· | ···················· |
| Z | 2887 | hCoV-19/USA/CA-CZB-14240/2020\|EPI_ISL_739276\|2020- | ···················· | ···················· | ····················· | ···················· | ···················· | ···················· |
| Z | 2888 | hCoV-19/USA/CA-CZB-15217/2020\|EPI_ISL_739277\|2020- | ···················· | ···················· | ····················· | ···················· | ···················· | ···················· |
| Z | 2890 | hCoV-19/USA/CA-CZB-14905/2020\|EPI_ISL_739279\|2020- | ···················· | ·A·················· | ····················· | ···················· | ···················· | ···················· |
| Z | 2891 | hCoV-19/USA/CA-CZB-13823/2020\|EPI_ISL_739280\|2020- | ···················· | ·A·················· | ····················· | ···················· | ···················· | ···················· |
| Z | 2892 | hCoV-19/USA/CA-CZB-14571/2020\|EPI_ISL_739281\|2020- | ···················· | ·A·················· | ····················· | ···················· | ···················· | ···················· |
| Z | 2893 | hCoV-19/USA/CA-CZB-15312/2020\|EPI_ISL_739282\|2020- | ···················· | ·A·················· | ····················· | ···················· | ···················· | ···················· |
| Z | 2895 | hCoV-19/USA/CA-CZB-14959/2020\|EPI_ISL_739284\|2020- | ···················· | ·A·················· | ····················· | ···················· | ···················· | ···················· |
| Z | 2896 | hCoV-19/USA/CA-CZB-13793/2020\|EPI_ISL_739285\|2020- | ···················· | ·A·················· | ····················· | ···················· | ···················· | ···················· |
| Z | 2898 | hCoV-19/USA/CA-CZB-14808/2020\|EPI_ISL_739287\|2020- | ···················· | ···················· | ····················· | ···················· | ···················· | ···················· |
| Z | 2899 | hCoV-19/USA/CA-CZB-13735/2020\|EPI_ISL_739288\|2020- | ···················· | ·A·················· | ····················· | ···················· | ···················· | ···················· |
| Z | 2900 | hCoV-19/USA/CA-CZB-14698/2020\|EPI_ISL_739289\|2020- | ···················· | ···················· | ····················· | ···················· | ···················· | ···················· |
| Z | 2902 | hCoV-19/USA/CA-CZB-14214/2020\|EPI_ISL_739291\|2020- | ···················· | ·············A······ | ····················· | ···················· | ···················· | ···················· |
| Z | 2903 | hCoV-19/USA/CA-CZB-14762/2020\|EPI_ISL_739292\|2020- | ···················· | ···················· | ····················· | ···················· | ···················· | ···················· |
| Z | 2904 | hCoV-19/USA/CA-CZB-14572/2020\|EPI_ISL_739293\|2020- | ···················· | ···················· | ····················· | ···················· | ···················· | ···················· |
| Z | 2906 | hCoV-19/USA/CA-CZB-14596/2020\|EPI_ISL_739295\|2020- | ···················· | ·A·················· | ····················· | ···················· | ···················· | ···················· |
| Z | 2907 | hCoV-19/USA/CA-CZB-14647/2020\|EPI_ISL_739296\|2020- | ···················· | ···················· | ····················· | ···················· | ···················· | ···················· |
| Z | 2908 | hCoV-19/USA/CA-CZB-14727/2020\|EPI_ISL_739297\|2020- | ···················· | ·A·················· | ····················· | ···················· | ···················· | ···················· |
| Z | 2909 | hCoV-19/USA/CA-CZB-15965/2020\|EPI_ISL_739298\|2020- | ···················· | ·A·················· | ····················· | ···················· | ···················· | ···················· |
| Z | 2910 | hCoV-19/USA/CA-CZB-14841/2020\|EPI_ISL_739299\|2020- | ···················· | ···················· | ····················· | ···················· | ···················· | ···················· |
| Z | 2911 | hCoV-19/Luxembourg/LNS2833134/2020\|EPI_ISL_740200\| | ···················· | ···················· | ····················· | ···················· | ···················· | ···················· |
| Z | 2912 | hCoV-19/Luxembourg/LNS2840790/2020\|EPI_ISL_740201\| | ···················· | ···················· | ····················· | ···················· | ······T············· | ···················· |
| Z | 2913 | hCoV-19/Luxembourg/LNS2846662/2020\|EPI_ISL_740203\| | ···················· | ···················· | ····················· | ···················· | ···················· | ···················· |
| Z | 2914 | hCoV-19/Luxembourg/LNS2851525/2020\|EPI_ISL_740205\| | ···················· | ···················· | ····················· | ···················· | ···················· | ···················· |
| Z | 2915 | hCoV-19/Luxembourg/LNS2852282/2020\|EPI_ISL_740206\| | ···················· | ···················· | ····················· | ···················· | ···················· | ···················· |
| Z | 2916 | hCoV-19/Luxembourg/LNS2856175/2020\|EPI_ISL_740207\| | ···················· | ···················· | ····················· | ···················· | ···················· | ···················· |
| Z | 2917 | hCoV-19/Luxembourg/LNS2857303/2020\|EPI_ISL_740208\| | ···················· | ···················· | ····················· | ···················· | ···················· | ···················· |
| Z | 2918 | hCoV-19/Luxembourg/LNS2868208/2020\|EPI_ISL_740209\| | ···················· | ···················· | ····················· | ···················· | ···················· | ···················· |
| Z | 2919 | hCoV-19/Luxembourg/LNS2878387/2020\|EPI_ISL_740210\| | ···················· | ···················· | ····················· | ···················· | ···················· | ···················· |
| Z | 2920 | hCoV-19/Luxembourg/LNS2883728/2020\|EPI_ISL_740211\| | ···················· | ···················· | ····················· | ···················· | ······T············· | ···················· |
| Z | 2921 | hCoV-19/Luxembourg/LNS2884060/2020\|EPI_ISL_740212\| | ···················· | ···················· | ····················· | ···················· | ···················· | ···················· |
| Z | 2922 | hCoV-19/Luxembourg/LNS2891686/2020\|EPI_ISL_740213\| | ···················· | ···················· | ····················· | ···················· | ···················· | ···················· |
| Z | 2923 | hCoV-19/Luxembourg/LNS2918076/2020\|EPI_ISL_740218\| | ···················· | ···················· | ····················· | ···················· | ···················· | ···················· |
| Z | 2924 | hCoV-19/Luxembourg/LNS2960590/2020\|EPI_ISL_740225\| | ···················· | ···················· | ····················· | ···················· | ···················· | ···················· |
| Z | 2925 | hCoV-19/Luxembourg/LNS2961665/2020\|EPI_ISL_740226\| | ···················· | ···················· | ····················· | ···················· | ···················· | ···················· |
| Z | 2926 | hCoV-19/Luxembourg/LNS2964698/2020\|EPI_ISL_740227\| | ···················· | ···················· | ····················· | ···················· | ···················· | ···················· |
| Z | 2927 | hCoV-19/Luxembourg/LNS2965079/2020\|EPI_ISL_740228\| | ···················· | ···················· | ····················· | ···················· | ···················· | ···················· |
| Z | 2928 | hCoV-19/Luxembourg/LNS2971553/2020\|EPI_ISL_740229\| | ···················· | ···················· | ····················· | ···················· | ···················· | ···················· |
| Z | 2929 | hCoV-19/Luxembourg/LNS2974720/2020\|EPI_ISL_740230\| | ···················· | ···················· | ····················· | ···················· | ···················· | ···················· |
| Z | 2930 | hCoV-19/Luxembourg/LNS2982165/2020\|EPI_ISL_740231\| | ···················· | ···················· | ····················· | ···················· | ···················· | ···················· |

Z 2931 hCoV-19/Luxembourg/LNS2994623/2020|EPI_ISL_740232| ···················· ···················· ····················· ···················· ···················· ····················
Z 2932 hCoV-19/Luxembourg/LNS3016414/2020|EPI_ISL_740233| ···················· ···················· ····················· ···················· ···················· ····················
Z 2933 hCoV-19/Luxembourg/LNS3019031/2020|EPI_ISL_740234| ···················· ···················· ····················· ···················· ···················· ····················
Z 2934 hCoV-19/Luxembourg/LNS3029641/2020|EPI_ISL_740235| ···················· ···················· ····················· ···················· ···················· ····················
Z 2935 hCoV-19/Luxembourg/LNS3030407/2020|EPI_ISL_740236| ···················· ···················· ···········T········· ···················· ···················· ····················
Z 2936 hCoV-19/Luxembourg/LNS3036725/2020|EPI_ISL_740237| ···················· ···················· ····················· ···················· ···················· ····················
Z 2937 hCoV-19/Luxembourg/LNS3048095/2020|EPI_ISL_740238| ···················· ···················· ····················· ···················· ···················· ····················
Z 2938 hCoV-19/Luxembourg/LNS3053849/2020|EPI_ISL_740239| ···················· ···················· ····················· ···················· ······T············· ····················
Z 2939 hCoV-19/Luxembourg/LNS3055496/2020|EPI_ISL_740240| ···················· ···················· ····················· ···················· ···················· ····················
Z 2940 hCoV-19/Luxembourg/LNS3061009/2020|EPI_ISL_740241| ···················· ····A··············· ····················· ···················· ···················· ····················
Z 2941 hCoV-19/Luxembourg/LNS3063798/2020|EPI_ISL_740242| ···················· ···················· ····················· ···················· ···················· ····················
Z 2942 hCoV-19/Luxembourg/LNS3085713/2020|EPI_ISL_740243| ···················· ···················· ····················· ···················· ···················· ····················
Z 2943 hCoV-19/Luxembourg/LNS3088379/2020|EPI_ISL_740244| ···················· ···················· ····················· ···················· ···················· ····················
Z 2944 hCoV-19/Luxembourg/LNS3090194/2020|EPI_ISL_740245| ···················· ···················· ····················· ···················· ···················· ····················
Z 2945 hCoV-19/Luxembourg/LNS3108529/2020|EPI_ISL_740247| ···················· ···················· ····················· ···················· ···················· ····················
Z 2946 hCoV-19/Luxembourg/LNS3112898/2020|EPI_ISL_740248| ···················· ···················· ····················· ···················· ···················· ····················
Z 2947 hCoV-19/Luxembourg/LNS3113282/2020|EPI_ISL_740249| ···················· ·A·················· ····················· ···················· ···················· ····················
Z 2948 hCoV-19/Luxembourg/LNS3116383/2020|EPI_ISL_740251| ···················· ···················· ····················· ···················· ···················· ····················
Z 2949 hCoV-19/Luxembourg/LNS3126705/2020|EPI_ISL_740253| ···················· ···················· ····················· ···················· ···················· ····················
Z 2950 hCoV-19/Luxembourg/LNS3130831/2020|EPI_ISL_740254| ···················· ···················· ····················· ···················· ···················· ····················
Z 2951 hCoV-19/Luxembourg/LNS3139399/2020|EPI_ISL_740255| ···················· ···················· ····················· ···················· ···················· ····················
Z 2952 hCoV-19/Luxembourg/LNS3140674/2020|EPI_ISL_740256| ············T······· ···················· ····················· ···················· ···················· ····················
Z 2953 hCoV-19/Luxembourg/LNS3145817/2020|EPI_ISL_740257| ···················· ···················· ····················· ···················· ···················· ····················
Z 2954 hCoV-19/Luxembourg/LNS3154706/2020|EPI_ISL_740260| ···················· ···················· ····················· ···················· ···················· ····················
Z 2955 hCoV-19/Luxembourg/LNS3157930/2020|EPI_ISL_740261| ···················· ···················· ····················· ···················· ···················· ····················
Z 2956 hCoV-19/Luxembourg/LNS3178789/2020|EPI_ISL_740262| ···················· ···················· ····················· ···················· ···················· ····················
Z 2957 hCoV-19/Luxembourg/LNS3182490/2020|EPI_ISL_740263| ···················· ···················· ····················· ···················· ···················· ····················
Z 2958 hCoV-19/Luxembourg/LNS3185046/2020|EPI_ISL_740264| ···················· ···················· ····················· ···················· ···················· ····················
Z 2959 hCoV-19/Luxembourg/LNS3186627/2020|EPI_ISL_740265| ···················· ···················· ····················· ···················· ···················· ····················
Z 2960 hCoV-19/Luxembourg/LNS3192468/2020|EPI_ISL_740266| ···················· ···················· ····················· ···················· ···················· ····················
Z 2961 hCoV-19/Luxembourg/LNS3193283/2020|EPI_ISL_740267| ···················· ···················· ····················· ···················· ···················· ····················
Z 2962 hCoV-19/Luxembourg/LNS3193318/2020|EPI_ISL_740268| ···················· ···················· ····················· ···················· ······T············· ····················
Z 2963 hCoV-19/Luxembourg/LNS3199654/2020|EPI_ISL_740269| ···················· ···················· ····················· ···················· ···················· ····················
Z 2964 hCoV-19/Luxembourg/LNS3201351/2020|EPI_ISL_740270| ···················· ···················· ····················· ···················· ···················· ····················
Z 2965 hCoV-19/Luxembourg/LNS3202155/2020|EPI_ISL_740271| ···················· ···················· ····················· ···················· ···················· ····················
Z 2966 hCoV-19/Luxembourg/LNS3207409/2020|EPI_ISL_740272| ···················· ···················· ····················· ···················· ···················· ····················
Z 2967 hCoV-19/Luxembourg/LNS3211792/2020|EPI_ISL_740273| ···················· ···················· ····················· ···················· ···················· ····················
Z 2968 hCoV-19/Luxembourg/LNS3214350/2020|EPI_ISL_740274| ···················· ···················· ····················· ···················· ···················· ····················
Z 2969 hCoV-19/Luxembourg/LNS3218144/2020|EPI_ISL_740275| ···················· ···················· ····················· ···················· ···················· ····················
Z 2970 hCoV-19/Luxembourg/LNS3219758/2020|EPI_ISL_740276| ···················· ···················· ····················· ···················· ···················· ····················
Z 2971 hCoV-19/Luxembourg/LNS3224690/2020|EPI_ISL_740277| ···················· ···················· ····················· ···················· ···················· ····················
Z 2972 hCoV-19/Luxembourg/LNS3257852/2020|EPI_ISL_740279| ···················· ···················· ····················· ···················· ···················· ····················
Z 2973 hCoV-19/Luxembourg/LNS3265404/2020|EPI_ISL_740280| ···················· ···················· ····················· ···················· ···················· ····················
Z 2974 hCoV-19/Luxembourg/LNS3274577/2020|EPI_ISL_740281| ···················· ···················· ····················· ···················· ···················· ····················
Z 2975 hCoV-19/Luxembourg/LNS3275003/2020|EPI_ISL_740282| ···················· ···················· ····················· ···················· ···················· ····················
Z 2976 hCoV-19/Luxembourg/LNS3280585/2020|EPI_ISL_740283| ···················· ···················· ····················· ···················· ···················· ····················
Z 2977 hCoV-19/Luxembourg/LNS3281030/2020|EPI_ISL_740284| ···················· ···················· ····················· ···················· ···················· ····················
Z 2978 hCoV-19/Luxembourg/LNS3288217/2020|EPI_ISL_740285| ···················· ···················· ····················· ···················· ···················· ····················
Z 2979 hCoV-19/Luxembourg/LNS3289822/2020|EPI_ISL_740286| ···················· ···················· ····················· ···················· ···················· ····················
Z 2980 hCoV-19/Luxembourg/LNS3324466/2020|EPI_ISL_740288| ···················· ···················· ····················· ···················· ···················· ····················
Z 2981 hCoV-19/Luxembourg/LNS3325407/2020|EPI_ISL_740289| ···················· ···················· ····················· ···················· ···················· ····················
Z 2982 hCoV-19/Luxembourg/LNS3331781/2020|EPI_ISL_740290| ···················· ···················· ····················· ···················· ···················· ····················
Z 2983 hCoV-19/Luxembourg/LNS3332260/2020|EPI_ISL_740291| ············T······· ···················· ····················· ···················· ···················· ····················
Z 2984 hCoV-19/Luxembourg/LNS3343013/2020|EPI_ISL_740292| ···················· ···················· ····················· ···················· ······T············· ····················
Z 2985 hCoV-19/Luxembourg/LNS3343273/2020|EPI_ISL_740293| ···················· ···················· ····················· ···················· ···················· ····················
Z 2986 hCoV-19/Luxembourg/LNS3350864/2020|EPI_ISL_740294| ···················· ···················· ····················· ···················· ···················· ····················
Z 2987 hCoV-19/Luxembourg/LNS3352091/2020|EPI_ISL_740295| ···················· ···················· ····················· ···················· ···················· ····················
Z 2988 hCoV-19/Luxembourg/LNS3355829/2020|EPI_ISL_740296| ···················· ···················· ····················· ···················· ···················· ····················
Z 2989 hCoV-19/Luxembourg/LNS3360603/2020|EPI_ISL_740297| ···················· ···················· ····················· ···················· ···················· ····················
Z 2990 hCoV-19/Luxembourg/LNS3361290/2020|EPI_ISL_740298| ···················· ···················· ····················· ···················· ···················· ····················
Z 2991 hCoV-19/Luxembourg/LNS3364439/2020|EPI_ISL_740299| ···················· ···················· ····················· ···················· ···················· ····················
Z 2992 hCoV-19/England/NORW-222CA8/2020|EPI_ISL_741512|20 ···················· ···················· ····················· ···················· ···················· ····················
Z 2993 hCoV-19/England/NORW-222CB7/2020|EPI_ISL_741513|20 ···················· ···················· ····················· ···················· ···················· ····················
Z 2994 hCoV-19/England/NORW-222CD5/2020|EPI_ISL_741514|20 ···················· ···················· ····················· ···················· ···················· ····················
Z 2995 hCoV-19/England/NORW-222D0F/2020|EPI_ISL_741517|20 ···················· ···················· ····················· ···················· ···················· ····················
Z 2996 hCoV-19/England/NORW-222D1E/2020|EPI_ISL_741518|20 ···················· ···················· ····················· ···················· ···················· ····················
Z 2997 hCoV-19/England/NORW-222D2D/2020|EPI_ISL_741519|20 ···················· ···················· ····················· ···················· ···················· ····················
Z 2998 hCoV-19/England/NORW-222D3C/2020|EPI_ISL_741520|20 ···················· ···················· ····················· ···················· ···················· ····················
Z 2999 hCoV-19/England/NORW-222D4B/2020|EPI_ISL_741521|20 ···················· ···················· ····················· ···················· ···················· ····················
Z 3000 hCoV-19/England/NORW-222D78/2020|EPI_ISL_741524|20 ···················· ·A·················· ····················· ···················· ···················· ···A················
Z 3001 hCoV-19/England/NORW-222D87/2020|EPI_ISL_741525|20 ···················· ···················· ····················· ···················· ···················· ····················
Z 3002 hCoV-19/England/NORW-222D96/2020|EPI_ISL_741526|20 ···················· ···················· ····················· ···················· ···················· ····················
Z 3003 hCoV-19/England/NORW-222DC3/2020|EPI_ISL_741529|20 ···················· ···················· ····················· ···················· ···················· ····················
Z 3004 hCoV-19/England/NORW-222DD2/2020|EPI_ISL_741530|20 ···················· ···················· ····················· ···················· ···················· ····················
Z 3005 hCoV-19/England/NORW-222DE1/2020|EPI_ISL_741531|20 ···················· ···················· ····················· ···················· ···················· ····················
Z 3006 hCoV-19/England/NORW-222E0C/2020|EPI_ISL_741533|20 ···················· ···················· ····················· ···················· ···················· ····················
Z 3007 hCoV-19/England/NORW-222E1B/2020|EPI_ISL_741534|20 ···················· ···················· ····················· ···················· ···················· ····················
Z 3008 hCoV-19/England/NORW-222E2A/2020|EPI_ISL_741535|20 ···················· ···················· ····················· ···················· ···················· ····················
Z 3009 hCoV-19/England/NORW-222E39/2020|EPI_ISL_741536|20 ···················· ···················· ····················· ···················· ···················· ····················
Z 3010 hCoV-19/England/NORW-222E66/2020|EPI_ISL_741538|20 ···················· ···················· ····················· ···················· ···················· ····················

| | | | | | | | | | |
|---|---|---|---|---|---|---|---|---|---|
| Z | 3011 | hCoV-19/England/NORW-222E75/2020\|EPI_ISL_741539\|20 | .................... | .................... | .................... | .................... | .................... | .................... | |
| Z | 3012 | hCoV-19/England/NORW-222E84/2020\|EPI_ISL_741540\|20 | .................... | .................... | .................... | .................... | .................... | .................... | |
| Z | 3013 | hCoV-19/England/NORW-222E93/2020\|EPI_ISL_741541\|20 | .................... | .................... | .................... | .................... | .................... | .................... | |
| Z | 3014 | hCoV-19/England/NORW-222EA2/2020\|EPI_ISL_741542\|20 | .................... | .A.................. | .................... | .................... | .................... | ..A................. | |
| Z | 3015 | hCoV-19/England/NORW-222EB1/2020\|EPI_ISL_741543\|20 | .................... | .................... | .................... | .................... | .................... | .................... | |
| Z | 3016 | hCoV-19/England/NORW-222EC0/2020\|EPI_ISL_741544\|20 | .................... | .................... | .................... | .................... | .................... | .................... | |
| Z | 3017 | hCoV-19/England/NORW-22307E/2020\|EPI_ISL_741558\|20 | .................... | .................... | .................... | .................... | .................... | .................... | |
| Z | 3018 | hCoV-19/England/NORW-22322D/2020\|EPI_ISL_741574\|20 | .................... | .................... | .................... | .................... | .................... | .................... | |
| Z | 3019 | hCoV-19/England/NORW-22323C/2020\|EPI_ISL_741575\|20 | .................... | .................... | .................... | .................... | .................... | .................... | |
| Z | 3020 | hCoV-19/England/NORW-22324B/2020\|EPI_ISL_741576\|20 | .................... | .................... | .................... | .................... | .................... | .................... | |
| Z | 3021 | hCoV-19/England/NORW-22325A/2020\|EPI_ISL_741577\|20 | .................... | .................... | .................... | .................... | .................... | .................... | |
| Z | 3022 | hCoV-19/England/NORW-223287/2020\|EPI_ISL_741579\|20 | .................... | .................... | .................... | .................... | .................... | .................... | |
| Z | 3023 | hCoV-19/England/NORW-2232B4/2020\|EPI_ISL_741581\|20 | .................... | .................... | .................... | .................... | .................... | .................... | |
| Z | 3024 | hCoV-19/England/NORW-F6577/2020\|EPI_ISL_741586\|202 | .................... | .................... | .................... | .................... | .................... | .................... | |
| Z | 3025 | hCoV-19/England/NORW-F9A0C/2020\|EPI_ISL_741588\|202 | .................... | .A.................. | .................... | .................... | .................... | ..A................. | |
| Z | 3026 | hCoV-19/England/NORW-F9BFA/2020\|EPI_ISL_741589\|202 | .................... | .................... | .................... | .................... | .................... | .................... | |
| Z | 3027 | hCoV-19/England/NORW-F9CCA/2020\|EPI_ISL_741593\|202 | .................... | .................... | .................... | .................... | .................... | .................... | |
| Z | 3028 | hCoV-19/England/NORW-F9CD9/2020\|EPI_ISL_741594\|202 | .................... | .................... | .................... | .................... | .................... | .................... | |
| Z | 3029 | hCoV-19/England/NORW-F9D5E/2020\|EPI_ISL_741596\|202 | .................... | .A.................. | .................... | .................... | .................... | ..A................. | |
| Z | 3030 | hCoV-19/England/NORW-F9D9A/2020\|EPI_ISL_741598\|202 | .................... | .R.................. | .................... | .................... | .................... | ..R................. | |
| Z | 3031 | hCoV-19/Wales/PHWC-49D8B1/2020\|EPI_ISL_742843\|2020 | .................... | .................... | .................... | .................... | .................... | .................... | |
| Z | 3032 | hCoV-19/Wales/PHWC-49D8C0/2020\|EPI_ISL_742844\|2020 | .................... | .................... | .................... | .................... | .................... | .................... | |
| Z | 3033 | hCoV-19/Wales/PHWC-49D8DF/2020\|EPI_ISL_742845\|2020 | .................... | .................... | .................... | .................... | .................... | .................... | |
| Z | 3034 | hCoV-19/Wales/PHWC-49D8FD/2020\|EPI_ISL_742846\|2020 | .................... | .................... | .................... | .................... | .................... | .................... | |
| Z | 3035 | hCoV-19/Wales/PHWC-49D918/2020\|EPI_ISL_742848\|2020 | .................... | .................... | .................... | .................... | .................... | .................... | |
| Z | 3036 | hCoV-19/Wales/PHWC-49D927/2020\|EPI_ISL_742849\|2020 | .................... | .................... | .................... | .................... | .................... | .................... | |
| Z | 3037 | hCoV-19/Wales/PHWC-49D936/2020\|EPI_ISL_742850\|2020 | .................... | .................... | .................... | .................... | .................... | .................... | |
| Z | 3038 | hCoV-19/Wales/PHWC-49D954/2020\|EPI_ISL_742852\|2020 | .................... | .................... | .................... | .................... | .................... | .................... | |
| Z | 3040 | hCoV-19/Wales/PHWC-49D981/2020\|EPI_ISL_742855\|2020 | .................... | .................... | .................... | .................... | .................... | .................... | |
| Z | 3041 | hCoV-19/Wales/PHWC-49D9AF/2020\|EPI_ISL_742856\|2020 | .................... | .................... | .................... | .................... | .................... | .................... | |
| Z | 3042 | hCoV-19/Wales/PHWC-49D9BE/2020\|EPI_ISL_742857\|2020 | .................... | .................... | .................... | .................... | .................... | .................... | |
| Z | 3043 | hCoV-19/Wales/PHWC-49D9DC/2020\|EPI_ISL_742859\|2020 | .................... | .................... | .................... | .................... | .................... | .................... | |
| Z | 3044 | hCoV-19/Wales/PHWC-49D9EB/2020\|EPI_ISL_742860\|2020 | .................... | .................... | .................... | .................... | .................... | .................... | |
| Z | 3045 | hCoV-19/Wales/PHWC-49D9FA/2020\|EPI_ISL_742861\|2020 | .................... | .................... | .................... | .................... | .................... | .................... | |
| Z | 3046 | hCoV-19/Wales/PHWC-49DA06/2020\|EPI_ISL_742862\|2020 | .................... | .................... | .................... | .................... | .................... | .................... | |
| Z | 3048 | hCoV-19/Wales/PHWC-49DA33/2020\|EPI_ISL_742864\|2020 | .................... | .................... | .................... | .................... | .................... | .................... | |
| Z | 3049 | hCoV-19/Wales/PHWC-49DA60/2020\|EPI_ISL_742866\|2020 | .................... | .................... | .................... | .................... | .................... | .................... | |
| Z | 3050 | hCoV-19/Wales/PHWC-49DA9D/2020\|EPI_ISL_742869\|2020 | .................... | .................... | .................... | .................... | .................... | .................... | |
| Z | 3051 | hCoV-19/Wales/PHWC-49DAAC/2020\|EPI_ISL_742870\|2020 | .................... | .................... | .................... | .................... | .................... | .................... | |
| Z | 3052 | hCoV-19/Wales/PHWC-49DABB/2020\|EPI_ISL_742871\|2020 | .................... | .................... | .................... | .................... | .................... | .................... | |
| Z | 3053 | hCoV-19/Wales/PHWC-49DACA/2020\|EPI_ISL_742872\|2020 | .................... | .................... | .................... | .................... | .................... | .................... | |
| Z | 3054 | hCoV-19/Wales/PHWC-49DAE8/2020\|EPI_ISL_742874\|2020 | .................... | .................... | .................... | .................... | .................... | .................... | |
| Z | 3055 | hCoV-19/Wales/PHWC-49DB03/2020\|EPI_ISL_742875\|2020 | .................... | .................... | .................... | .................... | .................... | .................... | |
| Z | 3056 | hCoV-19/Wales/PHWC-49DC3D/2020\|EPI_ISL_742888\|2020 | .................... | .................... | .................... | .................... | .................... | .................... | |
| Z | 3057 | hCoV-19/Wales/PHWC-49DC6A/2020\|EPI_ISL_742891\|2020 | .................... | .................... | .................... | .................... | .................... | .................... | |
| Z | 3058 | hCoV-19/Wales/PHWC-49DC88/2020\|EPI_ISL_742892\|2020 | .................... | .................... | .................... | .................... | NNN................. | .................... | 1 |
| Z | 3060 | hCoV-19/Wales/PHWC-49DCB5/2020\|EPI_ISL_742895\|2020 | .................... | .................... | .................... | .................... | .................... | .................... | |
| Z | 3061 | hCoV-19/Wales/PHWC-49DCD3/2020\|EPI_ISL_742896\|2020 | .................... | .................... | .................... | .................... | .................... | .................... | |
| Z | 3063 | hCoV-19/USA/UT-UPHL-2012331699/2020\|EPI_ISL_746440 | .................... | .A.................. | .................... | .................... | .................... | .................... | |
| Z | 3064 | hCoV-19/USA/UT-UPHL-2012341015/2020\|EPI_ISL_746441 | .................... | .................... | .................... | .................... | .................... | .................... | |
| Z | 3065 | hCoV-19/USA/UT-UPHL-2012361621/2020\|EPI_ISL_746442 | .................... | .................... | .................... | .................... | .................... | .................... | |
| Z | 3066 | hCoV-19/USA/UT-UPHL-2012391352/2020\|EPI_ISL_746443 | .................... | .................... | .................... | .................... | .................... | .................... | |
| Z | 3067 | hCoV-19/USA/UT-UPHL-2012416649/2020\|EPI_ISL_746444 | .................... | .................... | .................... | .................... | .................... | .................... | |
| Z | 3068 | hCoV-19/USA/UT-UPHL-2012438254/2020\|EPI_ISL_746445 | .................... | .................... | .................... | .................... | .................... | .................... | |
| Z | 3069 | hCoV-19/USA/UT-UPHL-2012460530/2020\|EPI_ISL_746446 | .................... | ......A............. | .................... | .................... | .................... | .................... | |
| Z | 3070 | hCoV-19/USA/UT-UPHL-2012489717/2020\|EPI_ISL_746447 | .................... | .................... | .................... | .................... | .................... | .................... | |
| Z | 3071 | hCoV-19/USA/UT-UPHL-2012582417/2020\|EPI_ISL_746448 | .................... | .A.................. | .................... | .................... | .................... | .................... | |
| Z | 3072 | hCoV-19/USA/UT-UPHL-2012582762/2020\|EPI_ISL_746449 | .................... | .................... | .................... | .................... | .................... | .................... | |
| Z | 3073 | hCoV-19/USA/CA-CZB-13768/2020\|EPI_ISL_739300\|2020- | .................... | .................... | .................... | .................... | .................... | .................... | |
| Z | 3074 | hCoV-19/USA/CA-CZB-11261/2020\|EPI_ISL_739301\|2020- | .................... | .................... | .................... | .................... | .................... | .................... | |
| Z | 3077 | hCoV-19/USA/CA-CZB-14233/2020\|EPI_ISL_739304\|2020- | .................... | .................... | .................... | .................... | .................... | .................... | |
| Z | 3078 | hCoV-19/USA/CA-CZB-15065/2020\|EPI_ISL_739305\|2020- | .................... | .................... | .................... | .................... | .................... | .................... | |
| Z | 3079 | hCoV-19/USA/CA-CZB-14106/2020\|EPI_ISL_739306\|2020- | .................... | .................... | .................... | .................... | .................... | .................... | |
| Z | 3080 | hCoV-19/USA/CA-CZB-13969/2020\|EPI_ISL_739307\|2020- | .................... | .................... | .................... | .................... | .................... | .................... | |
| Z | 3081 | hCoV-19/USA/CA-CZB-13928/2020\|EPI_ISL_739308\|2020- | .................... | .................... | .................... | .................... | .................... | .................... | |
| Z | 3082 | hCoV-19/USA/CA-CZB-14061/2020\|EPI_ISL_739309\|2020- | .................... | .................... | .................... | .................... | .................... | .................... | |
| Z | 3083 | hCoV-19/USA/CA-CZB-15200/2020\|EPI_ISL_739310\|2020- | .................... | .A.................. | .................... | .................... | .................... | .................... | |
| Z | 3084 | hCoV-19/USA/CA-CZB-14736/2020\|EPI_ISL_739311\|2020- | .................... | .A.................. | ...............T.... | .................... | .................... | .................... | |
| Z | 3085 | hCoV-19/USA/CA-CZB-14269/2020\|EPI_ISL_739312\|2020- | .................... | .................... | .................... | .................... | .................... | .................... | |
| Z | 3086 | hCoV-19/USA/CA-CZB-14809/2020\|EPI_ISL_739313\|2020- | .................... | .................... | .................... | .................... | .................... | .................... | |
| Z | 3087 | hCoV-19/USA/CA-CZB-15456/2020\|EPI_ISL_739314\|2020- | .................... | .................... | .................... | .................... | .................... | .................... | |
| Z | 3088 | hCoV-19/USA/CA-CZB-14603/2020\|EPI_ISL_739315\|2020- | .................... | .................... | .................... | .................... | ..N................. | .................... | |
| Z | 3089 | hCoV-19/USA/CA-CZB-16058/2020\|EPI_ISL_739316\|2020- | .................... | .A.................. | .................... | .................... | .................... | .................... | |
| Z | 3091 | hCoV-19/USA/CA-CZB-14795/2020\|EPI_ISL_739318\|2020- | .................... | .................... | .................... | .................... | .................... | .................... | |
| Z | 3092 | hCoV-19/USA/CA-CZB-15974/2020\|EPI_ISL_739319\|2020- | .................... | .A.................. | .................... | .................... | .................... | .................... | |
| Z | 3093 | hCoV-19/USA/CA-CZB-14573/2020\|EPI_ISL_739335\|2020- | .................... | .A.................. | .................... | .................... | .................... | .................... | |
| Z | 3095 | hCoV-19/USA/CA-CZB-14335/2020\|EPI_ISL_739338\|2020- | .................... | .................... | .................... | .................... | .................... | .................... | |
| Z | 3096 | hCoV-19/USA/CA-CZB-11269/2020\|EPI_ISL_739339\|2020- | .................... | .................... | .................... | .................... | .................... | .................... | |
| Z | 3097 | hCoV-19/USA/CA-CZB-14394/2020\|EPI_ISL_739340\|2020- | .................... | .A.................. | .................... | .................... | .................... | .................... | |
| Z | 3098 | hCoV-19/USA/CA-CZB-15428/2020\|EPI_ISL_739341\|2020- | .................... | .................... | .................... | .................... | .................... | .................... | |

```
Z    3099 hCoV-19/USA/CA-CZB-15475/2020|EPI_ISL_739342|2020-    ····················  ····················  ···T·················  ····················  ····················  ····················
Z    3100 hCoV-19/USA/CA-CZB-15059/2020|EPI_ISL_739343|2020-    ····················  ····················  ·····················  ····················  ····················  ····················
Z    3101 hCoV-19/USA/CA-CZB-15948/2020|EPI_ISL_739344|2020-    ····················  ····················  ·····················  ····················  ····················  ····················
Z    3102 hCoV-19/USA/CA-CZB-14634/2020|EPI_ISL_739345|2020-    ····················  ····················  ·····················  ····················  ····················  ····················
Z    3103 hCoV-19/USA/CA-CZB-14587/2020|EPI_ISL_739346|2020-    ····················  ·A··················  ·····················  ····················  ····················  ····················
Z    3104 hCoV-19/USA/CA-CZB-14892/2020|EPI_ISL_739347|2020-    ····················  ····················  ·····················  ····················  ····················  ····················
Z    3105 hCoV-19/USA/CA-CZB-16177/2020|EPI_ISL_739348|2020-    ····················  ····················  ·····················  ····················  ····················  ····················
Z    3107 hCoV-19/USA/CA-CZB-14697/2020|EPI_ISL_739350|2020-    ····················  ····················  ·····················  ····················  ····················  ····················
Z    3108 hCoV-19/USA/CA-CZB-14423/2020|EPI_ISL_739351|2020-    ····················  ····················  ·····················  ····················  ····················  ····················
Z    3110 hCoV-19/USA/CA-CZB-14655/2020|EPI_ISL_739353|2020-    ····················  ····················  ·····················  ····················  ····················  ····················
Z    3111 hCoV-19/USA/CA-CZB-15986/2020|EPI_ISL_739354|2020-    ····················  ·A··················  ·····················  ····················  ····················  ····················
Z    3112 hCoV-19/USA/CA-CZB-16178/2020|EPI_ISL_739355|2020-    ····················  ····················  ·····················  ····················  ····················  ····················
Z    3115 hCoV-19/USA/CA-CZB-13944/2020|EPI_ISL_739358|2020-    ····················  ····················  ·····················  ····················  ····················  ····················
Z    3116 hCoV-19/USA/CA-CZB-15452/2020|EPI_ISL_739359|2020-    ····················  ····················  ·····················  ····················  ····················  ····················
Z    3117 hCoV-19/Luxembourg/LNS3367744/2020|EPI_ISL_740300|    ····················  ····················  ·····················  ····················  ····················  ····················
Z    3118 hCoV-19/Luxembourg/LNS3388064/2020|EPI_ISL_740301|    ····················  ····················  ·····················  ····················  ····················  ····················
Z    3119 hCoV-19/Luxembourg/LNS3497064/2020|EPI_ISL_740320|    ····················  ····················  ·····················  ····················  ····················  ····················
Z    3120 hCoV-19/Luxembourg/LNS3500241/2020|EPI_ISL_740322|    ····················  ····················  ·····················  ····················  ····················  ····················
Z    3121 hCoV-19/Luxembourg/LNS3515812/2020|EPI_ISL_740324|    ····················  ····················  ·····················  ····················  ······T·············  ····················
Z    3122 hCoV-19/Luxembourg/LNS3518731/2020|EPI_ISL_740325|    ····················  ····················  ·····················  ····················  ····················  ····················
Z    3123 hCoV-19/Luxembourg/LNS3523220/2020|EPI_ISL_740326|    ····················  ····················  ·····················  ····················  ····················  ····················
Z    3124 hCoV-19/Luxembourg/LNS3528637/2020|EPI_ISL_740327|    ····················  ····················  ·····················  ····················  ····················  ····················
Z    3125 hCoV-19/Luxembourg/LNS3529687/2020|EPI_ISL_740328|    ····················  ····················  ·····················  ····················  ····················  ····················
Z    3126 hCoV-19/Luxembourg/LNS3533793/2020|EPI_ISL_740329|    ····················  ····················  ·····················  ····················  ····················  ····················
Z    3127 hCoV-19/Luxembourg/LNS3541605/2020|EPI_ISL_740330|    ····················  ····················  ·····················  ····················  ····················  ····················
Z    3128 hCoV-19/Luxembourg/LNS3548031/2020|EPI_ISL_740331|    ····················  ····················  ·····················  ····················  ····················  ····················
Z    3129 hCoV-19/Luxembourg/LNS3550762/2020|EPI_ISL_740332|    ····················  ····················  ·····················  ····················  ····················  ····················
Z    3130 hCoV-19/Luxembourg/LNS3562449/2020|EPI_ISL_740333|    ····················  ····················  ·····················  ····················  ····················  ····················
Z    3131 hCoV-19/Luxembourg/LNS3566996/2020|EPI_ISL_740334|    ············T·······  ····················  ·····················  ····················  ····················  ····················
Z    3132 hCoV-19/Luxembourg/LNS3573004/2020|EPI_ISL_740335|    ····················  ····················  ·····················  ····················  ····················  ····················
Z    3133 hCoV-19/Luxembourg/LNS3577548/2020|EPI_ISL_740336|    ····················  ····················  ·····················  ····················  ····················  ····················
Z    3134 hCoV-19/Luxembourg/LNS3580243/2020|EPI_ISL_740337|    ····················  ····················  ·····················  ····················  ······T·············  ····················
Z    3135 hCoV-19/Luxembourg/LNS3584304/2020|EPI_ISL_740338|    ····················  G···················  ·····················  ····················  ····················  ····················
Z    3136 hCoV-19/Luxembourg/LNS3585525/2020|EPI_ISL_740339|    ····················  ····················  ·····················  ····················  ····················  ····················
Z    3137 hCoV-19/Luxembourg/LNS3591441/2020|EPI_ISL_740340|    ····················  ····················  ·····················  ····················  ····················  ····················
Z    3138 hCoV-19/Luxembourg/LNS3600994/2020|EPI_ISL_740341|    ····················  ····················  ·····················  ····················  ····················  ····················
Z    3139 hCoV-19/Luxembourg/LNS3609481/2020|EPI_ISL_740342|    ····················  ····················  ·····················  ····················  ····················  ····················
Z    3140 hCoV-19/Luxembourg/LNS3624141/2020|EPI_ISL_740343|    ····················  ····················  ·····················  ····················  ····················  ····················
Z    3141 hCoV-19/Luxembourg/LNS3630121/2020|EPI_ISL_740344|    ····················  ····················  ·····················  ····················  ····················  ····················
Z    3142 hCoV-19/Luxembourg/LNS3640938/2020|EPI_ISL_740345|    ····················  ····················  ·····················  ····················  ····················  ····················
Z    3143 hCoV-19/England/NORW-F9E00/2020|EPI_ISL_741600|202    ····················  ·A··················  ·····················  ····················  ····················  ····················
Z    3144 hCoV-19/England/NORW-F9E3D/2020|EPI_ISL_741601|202    ····················  ····················  ·····················  ····················  ····················  ····················
Z    3145 hCoV-19/England/NORW-F9E5B/2020|EPI_ISL_741602|202    ····················  ·A··················  ·····················  ····················  ····················  ···A················
Z    3146 hCoV-19/England/NORW-F9F94/2020|EPI_ISL_741604|202    ····················  ····················  ·····················  ····················  ····················  ····················
Z    3147 hCoV-19/England/NORW-F9FB2/2020|EPI_ISL_741605|202    ····················  ·A··················  ·····················  ····················  ····················  ···A················
Z    3148 hCoV-19/England/NORW-F9FC1/2020|EPI_ISL_741606|202    ····················  ····················  ·····················  ····················  ····················  ····················
Z    3149 hCoV-19/England/NORW-F9FD0/2020|EPI_ISL_741607|202    ····················  ····················  ·····················  ····················  ····················  ····················
Z    3150 hCoV-19/England/NORW-F9FEF/2020|EPI_ISL_741608|202    ····················  ····················  ·····················  ····················  ····················  ····················
Z    3151 hCoV-19/England/NOTT-119DB1/2020|EPI_ISL_741609|20    ····················  ····················  ·····················  ····················  ····················  ····················
Z    3152 hCoV-19/England/NOTT-11EA2E/2020|EPI_ISL_741611|20    ····················  ····················  ·····················  ····················  ····················  ····················
Z    3153 hCoV-19/England/NOTT-11EE04/2020|EPI_ISL_741612|20    ····················  ····················  ·····················  ····················  ····················  ····················
Z    3154 hCoV-19/England/NOTT-1209A4/2020|EPI_ISL_741614|20    ····················  ····················  ·····················  ····················  ····················  ····················
Z    3155 hCoV-19/England/NOTT-120ED2/2020|EPI_ISL_741619|20    ····················  ····················  ·····················  ····················  ····················  ····················
Z    3156 hCoV-19/England/NOTT-120F93/2020|EPI_ISL_741621|20    ····················  ····················  ·····················  ····················  ····················  ····················
Z    3157 hCoV-19/England/NOTT-120FC0/2020|EPI_ISL_741622|20    ····················  ····················  ·····················  ····················  ····················  ····················
Z    3158 hCoV-19/England/NOTT-121017/2020|EPI_ISL_741623|20    ····················  ····················  ·····················  ····················  ····················  ····················
Z    3159 hCoV-19/England/NOTT-121123/2020|EPI_ISL_741626|20    ····················  ····················  ·····················  ····················  ····················  ····················
Z    3160 hCoV-19/England/NOTT-12127B/2020|EPI_ISL_741627|20    ····················  ····················  ·····················  ····················  ····················  ····················
Z    3161 hCoV-19/England/NOTT-121299/2020|EPI_ISL_741629|20    ····················  ····················  ·····················  ····················  ····················  ····················
Z    3162 hCoV-19/England/NOTT-1212C6/2020|EPI_ISL_741630|20    ····················  ····················  ·····················  ····················  ····················  ····················
Z    3163 hCoV-19/England/NOTT-121387/2020|EPI_ISL_741631|20    ····················  ····················  ·····················  ····················  ····················  ····················
Z    3164 hCoV-19/England/NOTT-12141B/2020|EPI_ISL_741632|20    ····················  ····················  ·····················  ····················  ····················  ····················
Z    3165 hCoV-19/England/NOTT-1215FA/2020|EPI_ISL_741644|20    ····················  ····················  ·····················  ····················  ····················  ····················
Z    3166 hCoV-19/England/NOTT-121719/2020|EPI_ISL_741650|20    ····················  ····················  ·····················  ····················  ····················  ····················
Z    3167 hCoV-19/England/NOTT-121729/2020|EPI_ISL_741651|20    ····················  ····················  ·····················  ····················  ····················  ····················
Z    3168 hCoV-19/England/NOTT-121792/2020|EPI_ISL_741656|20    ····················  ····················  ·····················  ····················  ····················  ····················
Z    3169 hCoV-19/England/PORT-2D4E09/2020|EPI_ISL_741658|20    ····················  ····················  ·····················  ····················  ····················  ····················
Z    3170 hCoV-19/England/PORT-2D4E27/2020|EPI_ISL_741659|20    ····················  ····················  ·····················  ····················  ····················  ····················
Z    3171 hCoV-19/England/PORT-2D4FE8/2020|EPI_ISL_741672|20    ····················  ····················  ·····················  ····················  ····················  ····················
Z    3172 hCoV-19/England/PORT-2D503F/2020|EPI_ISL_741675|20    ····················  ·A··················  ·····················  ····················  ····················  ···A················
Z    3173 hCoV-19/Wales/PHWC-49DD1C/2020|EPI_ISL_742900|2020    ····················  ····················  ·····················  ····················  ····················  ····················
Z    3174 hCoV-19/Wales/PHWC-49DD2B/2020|EPI_ISL_742901|2020    ····················  ····················  ·····················  ····················  ····················  ····················
Z    3175 hCoV-19/Wales/PHWC-49DD3A/2020|EPI_ISL_742902|2020    ····················  ····················  ·····················  ····················  ····················  ····················
Z    3177 hCoV-19/Wales/PHWC-49DD58/2020|EPI_ISL_742904|2020    ····················  ····················  ·····················  ····················  ····················  ····················
Z    3178 hCoV-19/Wales/PHWC-49DD67/2020|EPI_ISL_742905|2020    ····················  ····················  ·····················  ····················  ····················  ····················
Z    3179 hCoV-19/Wales/PHWC-49DD85/2020|EPI_ISL_742907|2020    ····················  ····················  ·····················  ····················  ····················  ····················
Z    3180 hCoV-19/Wales/PHWC-49DD94/2020|EPI_ISL_742908|2020    ····················  ····················  ·····················  ····················  ····················  ····················
Z    3181 hCoV-19/Wales/PHWC-49DDA3/2020|EPI_ISL_742909|2020    ····················  ····················  ·····················  ····················  ····················  ····················
Z    3182 hCoV-19/Wales/PHWC-49DDB2/2020|EPI_ISL_742910|2020    ····················  ····················  ·····················  ····················  ····················  ····················
Z    3183 hCoV-19/Wales/PHWC-49DDC1/2020|EPI_ISL_742911|2020    ····················  ····················  ·····················  ····················  ····················  ····················
```

| | | | | | | | | |
|---|---|---|---|---|---|---|---|---|
| Z | 3184 | hCoV-19/Wales/PHWC-49DDD0/2020\|EPI_ISL_742912\|2020 | ···················· | ···················· | ···················· | ···················· | ···················· | ···················· |
| Z | 3185 | hCoV-19/Wales/PHWC-49DDEF/2020\|EPI_ISL_742913\|2020 | ···················· | ···················· | ···················· | ···················· | ···················· | ···················· |
| Z | 3186 | hCoV-19/Wales/PHWC-49DDFE/2020\|EPI_ISL_742914\|2020 | ···················· | ···················· | ···················· | ···················· | ···················· | ···················· |
| Z | 3188 | hCoV-19/Wales/PHWC-49DE19/2020\|EPI_ISL_742916\|2020 | ···················· | ···················· | ···················· | ···················· | ···················· | ···················· |
| Z | 3189 | hCoV-19/Wales/PHWC-49DE28/2020\|EPI_ISL_742917\|2020 | ···················· | ···················· | ···················· | ···················· | ···················· | ···················· |
| Z | 3190 | hCoV-19/Wales/PHWC-49DE46/2020\|EPI_ISL_742918\|2020 | ···················· | ···················· | ···················· | ···················· | ···················· | ···················· |
| Z | 3191 | hCoV-19/Wales/PHWC-49DE55/2020\|EPI_ISL_742919\|2020 | ···················· | ···················· | ···················· | ···················· | ···················· | ···················· |
| Z | 3192 | hCoV-19/Wales/PHWC-49DE64/2020\|EPI_ISL_742920\|2020 | ···················· | ···················· | ···················· | ···················· | ···················· | ···················· |
| Z | 3193 | hCoV-19/Wales/PHWC-49DE73/2020\|EPI_ISL_742921\|2020 | ···················· | ···················· | ···················· | ···················· | ···················· | ···················· |
| Z | 3194 | hCoV-19/Wales/PHWC-49DE82/2020\|EPI_ISL_742922\|2020 | ···················· | ······A············· | ···················· | ···················· | ···················· | ···················· |
| Z | 3195 | hCoV-19/Wales/PHWC-49DE91/2020\|EPI_ISL_742923\|2020 | ···················· | ···················· | ···················· | ···················· | ···················· | ···················· |
| Z | 3196 | hCoV-19/Wales/PHWC-49DEA0/2020\|EPI_ISL_742924\|2020 | ···················· | ···················· | ···················· | ···················· | ···················· | ···················· |
| Z | 3197 | hCoV-19/Wales/PHWC-49DEBF/2020\|EPI_ISL_742925\|2020 | ···················· | ···················· | ···················· | ···················· | ···················· | ···················· |
| Z | 3198 | hCoV-19/Wales/PHWC-49DECE/2020\|EPI_ISL_742926\|2020 | ···················· | ···················· | ···················· | ···················· | ···················· | ···················· |
| Z | 3199 | hCoV-19/Wales/PHWC-49DEDD/2020\|EPI_ISL_742927\|2020 | ···················· | ···················· | ···················· | ···················· | ···················· | ···················· |
| Z | 3200 | hCoV-19/Wales/PHWC-49DEFB/2020\|EPI_ISL_742928\|2020 | ···················· | ···················· | ···················· | ···················· | ···················· | ···················· |
| Z | 3201 | hCoV-19/Wales/PHWC-49DF07/2020\|EPI_ISL_742929\|2020 | ···················· | ···················· | ···················· | ···················· | ···················· | ···················· |
| Z | 3202 | hCoV-19/Wales/PHWC-49DF16/2020\|EPI_ISL_742930\|2020 | ···················· | ···················· | ···················· | ···················· | ···················· | ···················· |
| Z | 3203 | hCoV-19/Wales/PHWC-49DF25/2020\|EPI_ISL_742931\|2020 | ···················· | ···················· | ···················· | ···················· | ···················· | ···················· |
| Z | 3204 | hCoV-19/Wales/PHWC-49DF34/2020\|EPI_ISL_742932\|2020 | ···················· | ···················· | ···················· | ···················· | ···················· | ···················· |
| Z | 3205 | hCoV-19/Wales/PHWC-49DF43/2020\|EPI_ISL_742933\|2020 | ···················· | ···················· | ···················· | ···················· | ···················· | ···················· |
| Z | 3206 | hCoV-19/Wales/PHWC-49DF52/2020\|EPI_ISL_742934\|2020 | ···················· | ···················· | ···················· | ···················· | ···················· | ···················· |
| Z | 3207 | hCoV-19/Wales/PHWC-49DF61/2020\|EPI_ISL_742935\|2020 | ···················· | ···················· | ···················· | ···················· | ···················· | ···················· |
| Z | 3208 | hCoV-19/Wales/PHWC-49DF70/2020\|EPI_ISL_742936\|2020 | ···················· | ···················· | ···················· | ···················· | ···················· | ···················· |
| Z | 3209 | hCoV-19/Wales/PHWC-49DF8F/2020\|EPI_ISL_742937\|2020 | ···················· | ···················· | ···················· | ···················· | ···················· | ···················· |
| Z | 3210 | hCoV-19/Wales/PHWC-49DFCB/2020\|EPI_ISL_742938\|2020 | ···················· | ···················· | ···················· | ···················· | ···················· | ···················· |
| Z | 3211 | hCoV-19/Wales/PHWC-49DFDA/2020\|EPI_ISL_742939\|2020 | ···················· | ···················· | ···················· | ···················· | ···················· | ···················· |
| Z | 3212 | hCoV-19/Wales/PHWC-49DFE9/2020\|EPI_ISL_742940\|2020 | ···················· | ···················· | ···················· | ···················· | ···················· | ···················· |
| Z | 3213 | hCoV-19/Wales/PHWC-49DFF8/2020\|EPI_ISL_742941\|2020 | ···················· | ···················· | ···················· | ···················· | ···················· | ···················· |
| Z | 3214 | hCoV-19/Wales/PHWC-49E003/2020\|EPI_ISL_742942\|2020 | ···················· | ···················· | ···················· | ···················· | ···················· | ···················· |
| Z | 3215 | hCoV-19/Wales/PHWC-49E012/2020\|EPI_ISL_742943\|2020 | ···················· | ···················· | ···················· | ···················· | ···················· | ···················· |
| Z | 3216 | hCoV-19/Wales/PHWC-49E021/2020\|EPI_ISL_742944\|2020 | ···················· | ···················· | ···················· | ···················· | ···················· | ···················· |
| Z | 3217 | hCoV-19/Wales/PHWC-49E030/2020\|EPI_ISL_742945\|2020 | ···················· | ···················· | ···················· | ···················· | ···················· | ···················· |
| Z | 3218 | hCoV-19/Wales/PHWC-49E05E/2020\|EPI_ISL_742946\|2020 | ···················· | ···················· | ···················· | ···················· | ···················· | ···················· |
| Z | 3219 | hCoV-19/Wales/PHWC-49E06D/2020\|EPI_ISL_742947\|2020 | ···················· | ···················· | ···················· | ···················· | ···················· | ···················· |
| Z | 3220 | hCoV-19/Wales/PHWC-49E07C/2020\|EPI_ISL_742948\|2020 | ···················· | ···················· | ···················· | ···················· | ···················· | ···················· |
| Z | 3221 | hCoV-19/Wales/PHWC-49E09A/2020\|EPI_ISL_742949\|2020 | ···················· | ···················· | ···················· | ···················· | ···················· | ···················· |
| Z | 3222 | hCoV-19/Wales/PHWC-49E0A9/2020\|EPI_ISL_742950\|2020 | ···················· | ······A············· | ···················· | ···················· | ···················· | ···················· |
| Z | 3223 | hCoV-19/Wales/PHWC-49E0B8/2020\|EPI_ISL_742951\|2020 | ···················· | ······A············· | ···················· | ···················· | ···················· | ···················· |
| Z | 3224 | hCoV-19/Wales/PHWC-49E0C7/2020\|EPI_ISL_742952\|2020 | ···················· | ···················· | ···················· | ···················· | ···················· | ···················· |
| Z | 3225 | hCoV-19/Wales/PHWC-49E0E5/2020\|EPI_ISL_742953\|2020 | ···················· | ···················· | ···················· | ···················· | ···················· | ···················· |
| Z | 3226 | hCoV-19/Wales/PHWC-49E0F4/2020\|EPI_ISL_742954\|2020 | ···················· | ······A············· | ···················· | ···················· | ···················· | ···················· |
| Z | 3227 | hCoV-19/Wales/PHWC-49E100/2020\|EPI_ISL_742955\|2020 | ···················· | ···················· | ···················· | ···················· | ···················· | ···················· |
| Z | 3228 | hCoV-19/Wales/PHWC-49E11F/2020\|EPI_ISL_742956\|2020 | ···················· | ···················· | ···················· | ···················· | ···················· | ···················· |
| Z | 3229 | hCoV-19/Wales/PHWC-49E12E/2020\|EPI_ISL_742957\|2020 | ···················· | ···················· | ···················· | ···················· | ···················· | ···················· |
| Z | 3231 | hCoV-19/Wales/PHWC-49E14C/2020\|EPI_ISL_742959\|2020 | ···················· | ···················· | ···················· | ···················· | ···················· | ···················· |
| Z | 3232 | hCoV-19/Wales/PHWC-49E16A/2020\|EPI_ISL_742960\|2020 | ···················· | ···················· | ···················· | ···················· | ···················· | ···················· |
| Z | 3233 | hCoV-19/Wales/PHWC-49E179/2020\|EPI_ISL_742961\|2020 | ···················· | ···················· | ···················· | ···················· | ···················· | ···················· |
| Z | 3234 | hCoV-19/Wales/PHWC-49E188/2020\|EPI_ISL_742962\|2020 | ···················· | ···················· | ···················· | ···················· | ···················· | ···················· |
| Z | 3235 | hCoV-19/Wales/PHWC-49E197/2020\|EPI_ISL_742963\|2020 | ···················· | ···················· | ···················· | ···················· | ···················· | ···················· |
| Z | 3236 | hCoV-19/Wales/PHWC-49E276/2020\|EPI_ISL_742976\|2020 | ···················· | ···················· | ···················· | ···················· | ···················· | ···················· |
| Z | 3237 | hCoV-19/Wales/PHWC-49E2B2/2020\|EPI_ISL_742980\|2020 | ···················· | ···················· | ···················· | ···················· | ···················· | ···················· |
| Z | 3238 | hCoV-19/Wales/PHWC-49E2C1/2020\|EPI_ISL_742981\|2020 | ···················· | ···················· | ···················· | ···················· | ···················· | ···················· |
| Z | 3239 | hCoV-19/Wales/PHWC-49E2EF/2020\|EPI_ISL_742983\|2020 | ···················· | ···················· | ···················· | ···················· | ···················· | ···················· |
| Z | 3240 | hCoV-19/Wales/PHWC-49E2FE/2020\|EPI_ISL_742984\|2020 | ···················· | ···················· | ···················· | ···················· | ···················· | ···················· |
| Z | 3241 | hCoV-19/Wales/PHWC-49E30A/2020\|EPI_ISL_742985\|2020 | ···················· | ···················· | ···················· | ···················· | ···················· | ···················· |
| Z | 3242 | hCoV-19/Wales/PHWC-49E319/2020\|EPI_ISL_742986\|2020 | ···················· | ···················· | ···················· | ···················· | ···················· | ···················· |
| Z | 3243 | hCoV-19/Wales/PHWC-49E328/2020\|EPI_ISL_742987\|2020 | ···················· | ···················· | ···················· | ···················· | ···················· | ···················· |
| Z | 3245 | hCoV-19/Wales/PHWC-49E346/2020\|EPI_ISL_742989\|2020 | ···················· | ···················· | ···················· | ···················· | ···················· | ···················· |
| Z | 3246 | hCoV-19/Wales/PHWC-49E355/2020\|EPI_ISL_742990\|2020 | ···················· | ···················· | ···················· | ···················· | ···················· | ···················· |
| Z | 3247 | hCoV-19/Wales/PHWC-49E373/2020\|EPI_ISL_742992\|2020 | ···················· | ···················· | ···················· | ···················· | ···················· | ···················· |
| Z | 3248 | hCoV-19/Wales/PHWC-49E382/2020\|EPI_ISL_742993\|2020 | ···················· | ···················· | ···················· | ···················· | ···················· | ···················· |
| Z | 3249 | hCoV-19/Wales/PHWC-49E3A0/2020\|EPI_ISL_742994\|2020 | ···················· | ···················· | ···················· | ···················· | ···················· | ···················· |
| Z | 3250 | hCoV-19/Wales/PHWC-49E3BF/2020\|EPI_ISL_742995\|2020 | ···················· | ···················· | ···················· | ···················· | ···················· | ···················· |
| Z | 3251 | hCoV-19/Wales/PHWC-49E3CE/2020\|EPI_ISL_742996\|2020 | ···················· | ···················· | ···················· | ···················· | ···················· | ···················· |
| Z | 3252 | hCoV-19/Wales/PHWC-49E3DD/2020\|EPI_ISL_742997\|2020 | ···················· | ···················· | ···················· | ···················· | ···················· | ···················· |
| Z | 3253 | hCoV-19/Wales/PHWC-49E3EC/2020\|EPI_ISL_742998\|2020 | ···················· | ···················· | ···················· | ···················· | ···················· | ···················· |
| Z | 3254 | hCoV-19/Australia/NSW1269/2020\|EPI_ISL_745100\|2020 | ···················· | ···················· | ···················· | ···················· | ···················· | ···················· |
| Z | 3255 | hCoV-19/Australia/NSW1270/2020\|EPI_ISL_745101\|2020 | ···················· | ···················· | ···················· | ···················· | ···················· | ···················· |
| Z | 3256 | hCoV-19/Australia/NSW1271/2020\|EPI_ISL_745102\|2020 | ···················· | ···················· | ···················· | ···················· | ···················· | ···················· |
| Z | 3257 | hCoV-19/Australia/NSW1272/2020\|EPI_ISL_745103\|2020 | ···················· | ···················· | ···················· | ···················· | ···················· | ···················· |
| Z | 3258 | hCoV-19/Australia/NSW1273/2020\|EPI_ISL_745104\|2020 | ···················· | ···················· | ···················· | ···················· | ···················· | ···················· |
| Z | 3259 | hCoV-19/Australia/NSW1274/2020\|EPI_ISL_745105\|2020 | ···················· | ···················· | ···················· | ···················· | ···················· | ···················· |
| Z | 3260 | hCoV-19/Australia/NSW1282/2020\|EPI_ISL_745106\|2020 | ···················· | ···················· | ···················· | ···················· | ···················· | ···················· |
| Z | 3261 | hCoV-19/Australia/NSW1283/2020\|EPI_ISL_745107\|2020 | ···················· | ···················· | ···················· | ···················· | ···················· | ···················· |
| Z | 3262 | hCoV-19/Australia/NSW1277/2020\|EPI_ISL_745108\|2020 | ···················· | ···················· | ···················· | ···················· | ···················· | ···················· |
| Z | 3263 | hCoV-19/South Africa/Tygerberg-453/2020\|EPI_ISL_74 | ···················· | ·A·················· | ···················· | ···················· | ···················· | ···················· |
| Z | 3264 | hCoV-19/South Africa/Tygerberg-460/2020\|EPI_ISL_74 | ···················· | ···················· | ···················· | ···················· | ···················· | ···················· |
| Z | 3265 | hCoV-19/South Africa/Tygerberg-395/2020\|EPI_ISL_74 | ···················· | ·A·················· | ···················· | ···················· | ···················· | ···················· |
| Z | 3266 | hCoV-19/South Africa/Tygerberg-407/2020\|EPI_ISL_74 | ···················· | ···················· | ···················· | ···················· | ···················· | ···················· |

| | | | | | | | | |
|---|---|---|---|---|---|---|---|---|
| Z | 3267 | hCoV-19/South Africa/Tygerberg-397/2020\|EPI_ISL_74 | ···················· | ·A·················· | ···················· | ···················· | ···················· | ···················· |
| Z | 3268 | hCoV-19/South Africa/Tygerberg-432/2020\|EPI_ISL_74 | ···················· | ·A·················· | ···················· | ···················· | ···················· | ···················· |
| Z | 3269 | hCoV-19/India/TG-CCMB-BB568/2020\|EPI_ISL_745198\|20 | ···················· | ·A·················· | ···················· | ···················· | ···················· | ···A················ |
| Z | 3270 | hCoV-19/USA/UT-UPHL-2012438158/2020\|EPI_ISL_746424 | ···················· | ···················· | ···················· | ···················· | ···················· | ···················· |
| Z | 3271 | hCoV-19/USA/UT-UPHL-2012006109/2020\|EPI_ISL_746425 | ···················· | ···················· | ···················· | ···················· | ···················· | ···················· |
| Z | 3272 | hCoV-19/USA/UT-UPHL-2012127660/2020\|EPI_ISL_746426 | ···················· | ···················· | ···················· | ···················· | ···················· | ···················· |
| Z | 3273 | hCoV-19/USA/UT-UPHL-2012130465/2020\|EPI_ISL_746427 | ···················· | ···················· | ···················· | ···················· | ···················· | ···················· |
| Z | 3274 | hCoV-19/USA/UT-UPHL-2012141655/2020\|EPI_ISL_746428 | ···················· | ·A·················· | ···················· | ···················· | ···················· | ···················· |
| Z | 3275 | hCoV-19/USA/UT-UPHL-2012167605/2020\|EPI_ISL_746429 | ···················· | ···················· | ···················· | ···················· | ···················· | ···················· |
| Z | 3276 | hCoV-19/USA/UT-UPHL-2012174571/2020\|EPI_ISL_746430 | ···················· | ···················· | ···················· | ···················· | ···················· | ···················· |
| Z | 3277 | hCoV-19/USA/UT-UPHL-2012181172/2020\|EPI_ISL_746431 | ···················· | ···················· | ···················· | ···················· | ···················· | ···················· |
| Z | 3278 | hCoV-19/USA/UT-UPHL-2012191643/2020\|EPI_ISL_746432 | ···················· | ···················· | ···················· | ···················· | ···················· | ···················· |
| Z | 3279 | hCoV-19/USA/UT-UPHL-2012207614/2020\|EPI_ISL_746433 | ···················· | ···················· | ···················· | ···················· | ···················· | ···················· |
| Z | 3280 | hCoV-19/USA/UT-UPHL-2012222800/2020\|EPI_ISL_746434 | ···················· | ···················· | ···················· | ···················· | ···················· | ···················· |
| Z | 3281 | hCoV-19/USA/UT-UPHL-2012236648/2020\|EPI_ISL_746435 | ···················· | ···················· | ···················· | ···················· | ···················· | ···················· |
| Z | 3282 | hCoV-19/USA/UT-UPHL-2012268592/2020\|EPI_ISL_746436 | ···················· | ·A·················· | ···················· | ···················· | ···················· | ···················· |
| Z | 3283 | hCoV-19/USA/UT-UPHL-2012270416/2020\|EPI_ISL_746437 | ···················· | ·A·················· | ···················· | ···················· | ···················· | ···················· |
| Z | 3284 | hCoV-19/USA/UT-UPHL-2012275312/2020\|EPI_ISL_746438 | ···················· | ···················· | ···················· | ···················· | ···················· | ···················· |
| Z | 3285 | hCoV-19/USA/UT-UPHL-2012312153/2020\|EPI_ISL_746439 | ···················· | ···················· | ···················· | ···················· | ···················· | ···················· |
| Z | 3286 | hCoV-19/USA/UT-UPHL-2012595394/2020\|EPI_ISL_746450 | ···················· | ·A·················· | ···················· | ···················· | ···················· | ···················· |
| Z | 3287 | hCoV-19/USA/UT-UPHL-2012609081/2020\|EPI_ISL_746451 | ···················· | ···················· | ···················· | ···················· | ···················· | ···················· |
| Z | 3288 | hCoV-19/USA/UT-UPHL-2012614705/2020\|EPI_ISL_746452 | ···················· | ···················· | ···················· | ···················· | ···················· | ···················· |
| Z | 3289 | hCoV-19/USA/UT-UPHL-2012644535/2020\|EPI_ISL_746453 | ···················· | ···················· | ···················· | ···················· | ···················· | ···················· |
| Z | 3290 | hCoV-19/USA/UT-UPHL-2012665905/2020\|EPI_ISL_746454 | ···················· | ···················· | ···················· | ···················· | ···················· | ···················· |
| Z | 3291 | hCoV-19/USA/UT-UPHL-2012669491/2020\|EPI_ISL_746455 | ···················· | ·A·················· | ···················· | ···················· | ···················· | ···················· |
| Z | 3292 | hCoV-19/USA/UT-UPHL-2012673507/2020\|EPI_ISL_746456 | ···················· | ···················· | ···················· | ···················· | ···················· | ···················· |
| Z | 3293 | hCoV-19/USA/UT-UPHL-2012676129/2020\|EPI_ISL_746457 | ···················· | ···················· | ···················· | ···················· | ···················· | ···················· |
| Z | 3294 | hCoV-19/USA/UT-UPHL-2012683062/2020\|EPI_ISL_746458 | ···················· | ···················· | ···················· | ···················· | ···················· | ···················· |
| Z | 3295 | hCoV-19/USA/UT-UPHL-2012703708/2020\|EPI_ISL_746459 | ···················· | ···················· | ···················· | ···················· | ···················· | ···················· |
| Z | 3296 | hCoV-19/USA/UT-UPHL-2012704031/2020\|EPI_ISL_746460 | ···················· | ···················· | ···················· | ···················· | ···················· | ···················· |
| Z | 3297 | hCoV-19/USA/UT-UPHL-2012723027/2020\|EPI_ISL_746461 | ···················· | ·A·················· | ···················· | ···················· | ···················· | ···················· |
| Z | 3298 | hCoV-19/USA/UT-UPHL-2012743524/2020\|EPI_ISL_746462 | ···················· | ···················· | ···················· | ···················· | ···················· | ···················· |
| Z | 3299 | hCoV-19/USA/UT-UPHL-2012749027/2020\|EPI_ISL_746463 | ···················· | ·A·················· | ···················· | ···················· | ···················· | ···················· |
| Z | 3300 | hCoV-19/USA/UT-UPHL-2012749679/2020\|EPI_ISL_746464 | ···················· | ···················· | ···················· | ···················· | ···················· | ···················· |
| Z | 3301 | hCoV-19/USA/UT-UPHL-2012750540/2020\|EPI_ISL_746465 | ···················· | ···················· | ···················· | ···················· | ···················· | ···················· |
| Z | 3302 | hCoV-19/USA/UT-UPHL-2012768126/2020\|EPI_ISL_746466 | ···················· | ···················· | ···················· | ···················· | ···················· | ···················· |
| Z | 3303 | hCoV-19/USA/UT-UPHL-2012796558/2020\|EPI_ISL_746467 | ···················· | ···················· | ···················· | ···················· | ···················· | ···················· |
| Z | 3304 | hCoV-19/USA/UT-UPHL-2012825995/2020\|EPI_ISL_746468 | ···················· | ···················· | ···················· | ···················· | ···················· | ···················· |
| Z | 3305 | hCoV-19/USA/UT-UPHL-2012838843/2020\|EPI_ISL_746469 | ···················· | ·A·················· | ···················· | ···················· | ···················· | ···················· |
| Z | 3306 | hCoV-19/USA/UT-UPHL-2012849622/2020\|EPI_ISL_746470 | ···················· | ···················· | ···················· | ···················· | ···················· | ···················· |
| Z | 3307 | hCoV-19/USA/UT-UPHL-2012863648/2020\|EPI_ISL_746471 | ···················· | ···················· | ···················· | ···················· | ···················· | ···················· |
| Z | 3308 | hCoV-19/USA/UT-UPHL-2012904972/2020\|EPI_ISL_746472 | ···················· | ···················· | ···················· | ···················· | ···················· | ···················· |
| Z | 3309 | hCoV-19/USA/UT-UPHL-2012915062/2020\|EPI_ISL_746473 | ···················· | ···················· | ···················· | ···················· | ···················· | ···················· |
| Z | 3310 | hCoV-19/USA/UT-UPHL-2012922250/2020\|EPI_ISL_746474 | ···················· | ···················· | ···················· | ···················· | ···················· | ···················· |
| Z | 3311 | hCoV-19/USA/UT-UPHL-2012935315/2020\|EPI_ISL_746475 | ···················· | ···················· | ···················· | ···················· | ···················· | ···················· |
| Z | 3312 | hCoV-19/USA/UT-UPHL-2012949083/2020\|EPI_ISL_746476 | ···················· | ···················· | ···················· | ···················· | ···················· | ···················· |
| Z | 3313 | hCoV-19/USA/UT-UPHL-2012964498/2020\|EPI_ISL_746477 | ···················· | ···················· | ···················· | ···················· | ···················· | ···················· |
| Z | 3314 | hCoV-19/Chile/RM-168408/2020\|EPI_ISL_746478\|2020-0 | ···················· | ···················· | ···················· | ···················· | ···················· | ···················· |
| Z | 3315 | hCoV-19/Chile/NB-196349/2020\|EPI_ISL_746479\|2020-1 | ···················· | ···················· | ···················· | ···················· | ···················· | ···················· |
| Z | 3316 | hCoV-19/Chile/AR-192894/2020\|EPI_ISL_746480\|2020-0 | ···················· | ···················· | ···················· | ···················· | ···················· | ···················· |
| Z | 3317 | hCoV-19/Chile/AR-179001/2020\|EPI_ISL_746481\|2020-0 | ···················· | ···················· | ···················· | ···················· | ···················· | ···················· |
| Z | 3318 | hCoV-19/Chile/RM-82489/2020\|EPI_ISL_746482\|2020-05 | ···················· | ···················· | ···················· | ···················· | ···················· | ···················· |
| Z | 3319 | hCoV-19/Chile/RM-191993/2020\|EPI_ISL_746483\|2020-0 | ···················· | ···················· | ···················· | ···················· | ···················· | ···················· |
| Z | 3320 | hCoV-19/Chile/RM-88166-TC1/2020\|EPI_ISL_746484\|202 | ···················· | ···················· | ···················· | ···················· | ···················· | ···················· |
| Z | 3321 | hCoV-19/Chile/RM-153750/2020\|EPI_ISL_746486\|2020-0 | ···················· | ···················· | ···················· | ···················· | ···················· | ···················· |
| Z | 3322 | hCoV-19/Chile/RM-155625/2020\|EPI_ISL_746487\|2020-0 | ···················· | ···················· | ···················· | ···················· | ···················· | ···················· |
| Z | 3323 | hCoV-19/Chile/RM-155645/2020\|EPI_ISL_746488\|2020-0 | ···················· | ···················· | ···················· | ···················· | ···················· | ···················· |
| Z | 3324 | hCoV-19/Chile/RM-72436/2020\|EPI_ISL_746489\|2020-05 | ···················· | ···················· | ···················· | ···················· | ···················· | ···················· |
| Z | 3325 | hCoV-19/Chile/AN-195419/2020\|EPI_ISL_746492\|2020-1 | ···················· | ···················· | ···················· | ···················· | ···················· | ···············A···· |
| Z | 3326 | hCoV-19/Chile/RM-82462/2020\|EPI_ISL_746493\|2020-05 | ···················· | ···················· | ···················· | ···················· | ···················· | ···················· |
| Z | 3327 | hCoV-19/Chile/AN-195430/2020\|EPI_ISL_746498\|2020-1 | ···················· | ···················· | ···················· | ···················· | ···················· | ···················· |
| Z | 3328 | hCoV-19/Chile/AR-172172/2020\|EPI_ISL_746499\|2020-0 | ···················· | ···················· | ···················· | ···················· | ···················· | ···················· |
| Z | 3329 | hCoV-19/USA/CA-CZB-15271/2020\|EPI_ISL_739320\|2020- | ···················· | ···················· | ···················· | ···················· | ···················· | ···················· |
| Z | 3330 | hCoV-19/USA/CA-CZB-13702/2020\|EPI_ISL_739321\|2020- | ···················· | ···················· | ···················· | ···················· | ···················· | ···················· |
| Z | 3331 | hCoV-19/USA/CA-CZB-14903/2020\|EPI_ISL_739322\|2020- | ···················· | ···················· | ···················· | ···················· | ···················· | ···················· |
| Z | 3333 | hCoV-19/USA/CA-CZB-14383/2020\|EPI_ISL_739324\|2020- | ···················· | ···················· | ···················· | ···················· | ···················· | ···················· |
| Z | 3334 | hCoV-19/USA/CA-CZB-14604/2020\|EPI_ISL_739325\|2020- | ···················· | ···················· | ···················· | ···················· | ···················· | ···················· |
| Z | 3335 | hCoV-19/USA/CA-CZB-13856/2020\|EPI_ISL_739326\|2020- | ···················· | ···················· | ···················· | ···················· | ···················· | ···················· |
| Z | 3336 | hCoV-19/USA/CA-CZB-15005/2020\|EPI_ISL_739327\|2020- | ···················· | ·A·················· | ···················· | ···················· | ···················· | ···················· |
| Z | 3338 | hCoV-19/USA/CA-CZB-14953/2020\|EPI_ISL_739329\|2020- | ···················· | ···················· | ···················· | ···················· | ···················· | ···················· |
| Z | 3339 | hCoV-19/USA/CA-CZB-14777/2020\|EPI_ISL_739330\|2020- | ···················· | ···················· | ···················· | ···················· | ···················· | ···················· |
| Z | 3340 | hCoV-19/USA/CA-CZB-14235/2020\|EPI_ISL_739331\|2020- | ···················· | ···················· | ···················· | ···················· | ··N················· | ···················· |
| Z | 3341 | hCoV-19/USA/CA-CZB-14583/2020\|EPI_ISL_739332\|2020- | ···················· | ···················· | ···················· | ···················· | ···················· | ···················· |
| Z | 3342 | hCoV-19/USA/CA-CZB-14115/2020\|EPI_ISL_739333\|2020- | ···················· | ···················· | ···················· | ···················· | ···················· | ···················· |
| Z | 3343 | hCoV-19/USA/CA-CZB-14692/2020\|EPI_ISL_739334\|2020- | ···················· | ···················· | ···················· | ···················· | ···················· | ···················· |
| Z | 3344 | hCoV-19/USA/CA-CZB-14948/2020\|EPI_ISL_739336\|2020- | ···················· | ·A·················· | ···················· | ···················· | ···················· | ···················· |
| Z | 3345 | hCoV-19/USA/CA-CZB-15099/2020\|EPI_ISL_739360\|2020- | ···················· | ···················· | ···················· | ···················· | ···················· | ···················· |
| Z | 3346 | hCoV-19/USA/CA-CZB-13888/2020\|EPI_ISL_739361\|2020- | ···················· | ···················· | ···················· | ···················· | ···················· | ···················· |
| Z | 3347 | hCoV-19/USA/CA-CZB-14702/2020\|EPI_ISL_739362\|2020- | ···················· | ···················· | ···················· | ···················· | ···················· | ···················· |
| Z | 3348 | hCoV-19/USA/CA-CZB-15481/2020\|EPI_ISL_739363\|2020- | ···················· | ·A·················· | ···················· | ···················· | ···················· | ···················· |

Z 3349 hCoV-19/USA/CA-CZB-14952/2020|EPI_ISL_739364|2020- ···················· ···················· ····················· ···················· ···················· ····················
Z 3350 hCoV-19/USA/CA-CZB-15067/2020|EPI_ISL_739365|2020- ···················· ·A·················· ····················· ···················· ···················· ····················
Z 3351 hCoV-19/USA/CA-CZB-14730/2020|EPI_ISL_739366|2020- ···················· ·A·················· ···········T········· ···················· ···················· ····················
Z 3352 hCoV-19/USA/CA-CZB-14780/2020|EPI_ISL_739367|2020- ···················· ···················· ····················· ···················· ···················· ····················
Z 3353 hCoV-19/USA/CA-CZB-14311/2020|EPI_ISL_739368|2020- ···················· ·A·················· ····················· ···················· ···················· ····················
Z 3354 hCoV-19/USA/CA-CZB-13716/2020|EPI_ISL_739369|2020- ···················· ···················· ····················· ···················· ···················· ····················
Z 3355 hCoV-19/USA/CA-CZB-15977/2020|EPI_ISL_739370|2020- ···················· ·A·················· ····················· ···················· ···················· ····················
Z 3356 hCoV-19/USA/CA-CZB-15985/2020|EPI_ISL_739371|2020- ···················· ·A·················· ····················· ···················· ···················· ····················
Z 3357 hCoV-19/USA/CA-CZB-14950/2020|EPI_ISL_739372|2020- ···················· ·A·················· ····················· ···················· ···················· ····················
Z 3358 hCoV-19/USA/CA-CZB-14687/2020|EPI_ISL_739373|2020- ···················· ···················· ····················· ···················· ···················· ····················
Z 3359 hCoV-19/USA/CA-CZB-14565/2020|EPI_ISL_739374|2020- ···················· ···················· ····················· ···················· ···················· ····················
Z 3360 hCoV-19/USA/CA-CZB-13717/2020|EPI_ISL_739375|2020- ···················· ···················· ····················· ···················· ···················· ····················
Z 3362 hCoV-19/USA/CA-CZB-15092/2020|EPI_ISL_739377|2020- ···················· ···················· ····················· ···················· ···················· ····················
Z 3363 hCoV-19/USA/CA-CZB-14009/2020|EPI_ISL_739378|2020- ···················· ···················· ····················· ···················· ···················· ····················
Z 3364 hCoV-19/USA/CA-CZB-14239/2020|EPI_ISL_739379|2020- ···················· ···················· ····················· ···················· ···················· ····················
Z 3365 hCoV-19/USA/CA-CZB-15006/2020|EPI_ISL_739380|2020- ···················· ·A·················· ····················· ···················· ···················· ····················
Z 3366 hCoV-19/USA/CA-CZB-15945/2020|EPI_ISL_739381|2020- ···················· ···················· ····················· ···················· ···················· ····················
Z 3367 hCoV-19/USA/CA-CZB-15139/2020|EPI_ISL_739382|2020- ···················· ···················· ····················· ···················· ···················· ····················
Z 3368 hCoV-19/USA/CA-CZB-14145/2020|EPI_ISL_739383|2020- ···················· ···················· ····················· ···················· ···················· ····················
Z 3369 hCoV-19/USA/CA-CZB-13725/2020|EPI_ISL_739384|2020- ···················· ···················· ····················· ···················· ···················· ····················
Z 3370 hCoV-19/USA/CA-CZB-14010/2020|EPI_ISL_739385|2020- ···················· ···················· ····················· ···················· ···················· ····················
Z 3371 hCoV-19/USA/CA-CZB-14225/2020|EPI_ISL_739386|2020- ···················· ···················· ····················· ···················· ···················· ····················
Z 3372 hCoV-19/USA/CA-CZB-13870/2020|EPI_ISL_739387|2020- ···················· ···················· ····················· ···················· ···················· ····················
Z 3373 hCoV-19/USA/CA-CZB-13682/2020|EPI_ISL_739388|2020- ···················· ···················· ····················· ···················· ···················· ····················
Z 3374 hCoV-19/USA/CA-CZB-14797/2020|EPI_ISL_739389|2020- ···················· ···················· ····················· ···················· ···················· ····················
Z 3375 hCoV-19/USA/CA-CZB-14150/2020|EPI_ISL_739390|2020- ···················· ···················· ····················· ···················· ···················· ····················
Z 3376 hCoV-19/USA/CA-CZB-14404/2020|EPI_ISL_739391|2020- ···················· ···················· ····················· ···················· ···················· ····················
Z 3378 hCoV-19/USA/CA-CZB-14160/2020|EPI_ISL_739393|2020- ···················· ·A·················· ····················· ···················· ···················· ····················
Z 3379 hCoV-19/USA/CA-CZB-15994/2020|EPI_ISL_739394|2020- ···················· ·A·················· ····················· ···················· ···················· ····················
Z 3380 hCoV-19/USA/CA-CZB-14402/2020|EPI_ISL_739395|2020- ···················· ···A················ ····················· ···················· ···················· ····················
Z 3381 hCoV-19/USA/CA-CZB-13719/2020|EPI_ISL_739396|2020- ···················· ·A·················· ····················· ···················· ···················· ····················
Z 3382 hCoV-19/USA/CA-CZB-14648/2020|EPI_ISL_739397|2020- ···················· ···················· ····················· ···················· ···················· ····················
Z 3383 hCoV-19/USA/CA-CZB-15962/2020|EPI_ISL_739398|2020- ···················· ·A·················· ····················· ···················· ···················· ····················
Z 3384 hCoV-19/USA/CA-CZB-15497/2020|EPI_ISL_739399|2020- ···················· ···················· ····················· ···················· ···················· ····················
Z 3385 hCoV-19/Luxembourg/LNS3401997/2020|EPI_ISL_740303| ···················· ···················· ····················· ···················· ···················· ····················
Z 3386 hCoV-19/Luxembourg/LNS3413350/2020|EPI_ISL_740304| ···················· ···················· ····················· ···················· ···················· ····················
Z 3387 hCoV-19/Luxembourg/LNS3416856/2020|EPI_ISL_740305| ···················· ···················· ····················· ···················· ···················· ····················
Z 3388 hCoV-19/Luxembourg/LNS3417408/2020|EPI_ISL_740307| ···················· ···················· ····················· ···················· ···················· ····················
Z 3389 hCoV-19/Luxembourg/LNS3433209/2020|EPI_ISL_740309| ···················· ···················· ····················· ···················· ···················· ····················
Z 3390 hCoV-19/Luxembourg/LNS3438764/2020|EPI_ISL_740310| ···················· ···················· ····················· ···················· ···················· ····················
Z 3391 hCoV-19/Luxembourg/LNS3444661/2020|EPI_ISL_740311| ···················· ···················· ····················· ···················· ···················· ····················
Z 3392 hCoV-19/Luxembourg/LNS3459765/2020|EPI_ISL_740312| ···················· ···················· ····················· ···················· ···················· ····················
Z 3393 hCoV-19/Luxembourg/LNS3483404/2020|EPI_ISL_740314| ···················· ···················· ····················· ···················· ···················· ····················
Z 3394 hCoV-19/Luxembourg/LNS3485994/2020|EPI_ISL_740315| ···················· ···················· ····················· ···················· ······T············· ····················
Z 3395 hCoV-19/Luxembourg/LNS3488904/2020|EPI_ISL_740316| ···················· ···················· ····················· ···················· ···················· ····················
Z 3396 hCoV-19/Luxembourg/LNS3491081/2020|EPI_ISL_740317| ···················· ···················· ····················· ···················· ···················· ····················
Z 3397 hCoV-19/Luxembourg/LNS3493823/2020|EPI_ISL_740318| ···················· ···················· ····················· ···················· ···················· ····················
Z 3398 hCoV-19/Luxembourg/LNS3495560/2020|EPI_ISL_740319| ···················· ···················· ····················· ···················· ···················· ····················
Z 3399 hCoV-19/Luxembourg/LNS3647356/2020|EPI_ISL_740346| ···················· ···················· ····················· ···················· ···················· ····················
Z 3400 hCoV-19/Luxembourg/LNS3649868/2020|EPI_ISL_740347| ···················· ···················· ····················· ···················· ···················· ····················
Z 3401 hCoV-19/Luxembourg/LNS3652230/2020|EPI_ISL_740348| ············T······· ···················· ····················· ···················· ···················· ····················
Z 3402 hCoV-19/Luxembourg/LNS3654029/2020|EPI_ISL_740349| ···················· ···················· ····················· ···················· ···················· ····················
Z 3403 hCoV-19/Luxembourg/LNS3670718/2020|EPI_ISL_740350| ···················· ···················· ····················· ···················· ···················· ····················
Z 3404 hCoV-19/Luxembourg/LNS3670945/2020|EPI_ISL_740351| ···················· ···················· ····················· ···················· ···················· ····················
Z 3405 hCoV-19/Luxembourg/LNS3672765/2020|EPI_ISL_740352| ···················· ···················· ····················· ···················· ···················· ····················
Z 3406 hCoV-19/Luxembourg/LNS3677955/2020|EPI_ISL_740354| ···················· ···················· ····················· ···················· ···················· ····················
Z 3407 hCoV-19/Luxembourg/LNS3678271/2020|EPI_ISL_740355| ···················· ···················· ····················· ···················· ···················· ····················
Z 3408 hCoV-19/Luxembourg/LNS3679210/2020|EPI_ISL_740356| ···················· ···················· ····················· ···················· ···················· ····················
Z 3409 hCoV-19/Luxembourg/LNS3683612/2020|EPI_ISL_740357| ···················· ···················· ····················· ···················· ···················· ····················
Z 3410 hCoV-19/Luxembourg/LNS3688376/2020|EPI_ISL_740358| ···················· ···················· ····················· ···················· ···················· ····················
Z 3411 hCoV-19/Luxembourg/LNS3690941/2020|EPI_ISL_740359| ···················· ···················· ····················· ···················· ···················· ····················
Z 3412 hCoV-19/Luxembourg/LNS3693395/2020|EPI_ISL_740360| ···················· ···················· ····················· ···················· ···················· ····················
Z 3413 hCoV-19/Luxembourg/LNS3703401/2020|EPI_ISL_740361| ···················· ···················· ····················· ···················· ···················· ····················
Z 3414 hCoV-19/Luxembourg/LNS3705347/2020|EPI_ISL_740362| ···················· ···················· ····················· ···················· ······T············· ····················
Z 3415 hCoV-19/Luxembourg/LNS3708276/2020|EPI_ISL_740363| ···················· ···················· ····················· ···················· ···················· ····················
Z 3416 hCoV-19/Luxembourg/LNS3712604/2020|EPI_ISL_740364| ···················· ···················· ····················· ···················· ···················· ····················
Z 3417 hCoV-19/Luxembourg/LNS3723581/2020|EPI_ISL_740366| ···················· ···················· ····················· ···················· ···················· ····················
Z 3418 hCoV-19/Luxembourg/LNS3733630/2020|EPI_ISL_740367| ···················· ···················· ····················· ···················· ···················· ····················
Z 3419 hCoV-19/Luxembourg/LNS3741551/2020|EPI_ISL_740369| ···················· ···················· ····················· ···················· ···················· ····················
Z 3420 hCoV-19/Luxembourg/LNS3743453/2020|EPI_ISL_740370| ···················· ···················· ····················· ···················· ···················· ····················
Z 3421 hCoV-19/Luxembourg/LNS3758807/2020|EPI_ISL_740371| ···················· ···················· ····················· ···················· ···················· ····················
Z 3422 hCoV-19/Luxembourg/LNS3760280/2020|EPI_ISL_740372| ···················· ···················· ····················· ···················· ···················· ····················
Z 3423 hCoV-19/Luxembourg/LNS3775027/2020|EPI_ISL_740373| ···················· ···················· ····················· ···················· ···················· ····················
Z 3424 hCoV-19/Luxembourg/LNS3776268/2020|EPI_ISL_740374| ···················· ···················· ····················· ···················· ···················· ····················
Z 3425 hCoV-19/Luxembourg/LNS3778308/2020|EPI_ISL_740375| ···················· ···················· ····················· ···················· ···················· ····················
Z 3426 hCoV-19/Luxembourg/LNS3786534/2020|EPI_ISL_740376| ···················· ···················· ····················· ···················· ···················· ····················
Z 3427 hCoV-19/Luxembourg/LNS3827372/2020|EPI_ISL_740378| ···················· ···················· ····················· ···················· ···················· ····················
Z 3428 hCoV-19/Luxembourg/LNS3836811/2020|EPI_ISL_740379| ···················· ···················· ····················· ···················· ···················· ····················
Z 3429 hCoV-19/Luxembourg/LNS3838408/2020|EPI_ISL_740380| ···················· ···················· ····················· ···················· ···················· ····················
Z 3430 hCoV-19/Luxembourg/LNS3841144/2020|EPI_ISL_740381| ···················· ···················· ····················· ···················· ···················· ····················

| | | | | | | | |
|---|---|---|---|---|---|---|---|
| Z | 3431 hCoV-19/Luxembourg/LNS3841557/2020\|EPI_ISL_740382\| | ···········T········ | ···················· | ···················· | ···················· | ···················· | ···················· |
| Z | 3432 hCoV-19/Luxembourg/LNS3858275/2020\|EPI_ISL_740383\| | ···················· | ···················· | ···················· | ···················· | ······T············· | ···················· |
| Z | 3433 hCoV-19/Luxembourg/LNS3865010/2020\|EPI_ISL_740384\| | ···················· | ···················· | ···················· | ···················· | ···················· | ···················· |
| Z | 3434 hCoV-19/Luxembourg/LNS3865386/2020\|EPI_ISL_740385\| | ···················· | ···················· | ···················· | ···················· | ···················· | ···················· |
| Z | 3435 hCoV-19/Luxembourg/LNS3866946/2020\|EPI_ISL_740386\| | ···················· | ···················· | ···················· | ···················· | ···················· | ···················· |
| Z | 3436 hCoV-19/Luxembourg/LNS3877129/2020\|EPI_ISL_740387\| | ···················· | ···················· | ···················· | ···················· | ···················· | ···················· |
| Z | 3437 hCoV-19/Luxembourg/LNS3891670/2020\|EPI_ISL_740388\| | ···················· | ···················· | ···················· | ···················· | ···················· | ···················· |
| Z | 3438 hCoV-19/Luxembourg/LNS3920210/2020\|EPI_ISL_740389\| | ···················· | ···················· | ···················· | ···················· | ···················· | ···················· |
| Z | 3439 hCoV-19/Luxembourg/LNS3922198/2020\|EPI_ISL_740390\| | ···················· | ···················· | ···················· | ···················· | ···················· | ···················· |
| Z | 3440 hCoV-19/Luxembourg/LNS3922342/2020\|EPI_ISL_740391\| | ···················· | ···················· | ···················· | ···················· | ···················· | ···················· |
| Z | 3441 hCoV-19/Luxembourg/LNS3923367/2020\|EPI_ISL_740392\| | ···················· | ···················· | ···················· | ···················· | ···················· | ···················· |
| Z | 3442 hCoV-19/Luxembourg/LNS3931513/2020\|EPI_ISL_740393\| | ···················· | ···················· | ···················· | ···················· | ···················· | ···················· |
| Z | 3443 hCoV-19/Luxembourg/LNS3950198/2020\|EPI_ISL_740394\| | ···················· | ··G················· | ···················· | ···················· | ···················· | ···················· |
| Z | 3444 hCoV-19/Luxembourg/LNS3955853/2020\|EPI_ISL_740395\| | ···················· | ···················· | ···················· | ···················· | ···················· | ···················· |
| Z | 3445 hCoV-19/Luxembourg/LNS3959009/2020\|EPI_ISL_740396\| | ···················· | ···················· | ···················· | ···················· | ···················· | ···················· |
| Z | 3446 hCoV-19/Luxembourg/LNS3979799/2020\|EPI_ISL_740397\| | ···················· | ···················· | ···················· | ···················· | ······T············· | ···················· |
| Z | 3447 hCoV-19/Luxembourg/LNS3988953/2020\|EPI_ISL_740398\| | ···················· | ···················· | ···················· | ···················· | ···················· | ···················· |
| Z | 3448 hCoV-19/Luxembourg/LNS3989074/2020\|EPI_ISL_740399\| | ···················· | ···················· | ···················· | ···················· | ···················· | ···················· |
| Z | 3449 hCoV-19/England/NOTT-12142A/2020\|EPI_ISL_741633\|20 | ···················· | ···················· | ···················· | ···················· | ···················· | ···················· |
| Z | 3450 hCoV-19/England/NOTT-121439/2020\|EPI_ISL_741634\|20 | ···················· | ···················· | ···················· | ···················· | ···················· | ···················· |
| Z | 3451 hCoV-19/England/NOTT-121466/2020\|EPI_ISL_741637\|20 | ···················· | ···················· | ···················· | ···················· | ···················· | ···················· |
| Z | 3452 hCoV-19/England/NOTT-121475/2020\|EPI_ISL_741638\|20 | ···················· | ···················· | ···················· | ···················· | ···················· | ···················· |
| Z | 3453 hCoV-19/England/NOTT-121518/2020\|EPI_ISL_741639\|20 | ···················· | ···················· | ···················· | ···················· | ···················· | ···················· |
| Z | 3454 hCoV-19/England/NOTT-121554/2020\|EPI_ISL_741640\|20 | ···················· | ···················· | ···················· | ···················· | ···················· | ···················· |
| Z | 3455 hCoV-19/England/NOTT-121606/2020\|EPI_ISL_741645\|20 | ···················· | ···················· | ···················· | ···················· | ···················· | ···················· |
| Z | 3456 hCoV-19/England/NOTT-121707/2020\|EPI_ISL_741647\|20 | ···················· | ···················· | ···················· | ···················· | ···················· | ···················· |
| Z | 3457 hCoV-19/England/NOTT-121716/2020\|EPI_ISL_741649\|20 | ···················· | ···················· | ···················· | ···················· | ···················· | ···················· |
| Z | 3458 hCoV-19/England/PORT-2D513C/2020\|EPI_ISL_741680\|20 | ···················· | ···················· | ···················· | ···················· | ······T············· | ···················· |
| Z | 3459 hCoV-19/England/PORT-2D514B/2020\|EPI_ISL_741681\|20 | ···················· | ···················· | ···················· | ···················· | ······T············· | ···················· |
| Z | 3460 hCoV-19/England/PORT-2D515A/2020\|EPI_ISL_741682\|20 | ···················· | ···················· | ···················· | ···················· | ······T············· | ···················· |
| Z | 3461 hCoV-19/England/PORT-2D5178/2020\|EPI_ISL_741683\|20 | ···················· | ···················· | ···················· | ···················· | ······T············· | ···················· |
| Z | 3462 hCoV-19/England/PORT-2D5187/2020\|EPI_ISL_741684\|20 | ···················· | ···················· | ···················· | ···················· | ······T············· | ···················· |
| Z | 3463 hCoV-19/England/PORT-2D52EE/2020\|EPI_ISL_741694\|20 | ···················· | ···················· | ···················· | ···················· | ···················· | ···················· |
| Z | 3464 hCoV-19/Wales/PHWC-49E1A6/2020\|EPI_ISL_742964\|2020 | ···················· | ···················· | ···················· | ···················· | ···················· | ···················· |
| Z | 3465 hCoV-19/Wales/PHWC-49E1C4/2020\|EPI_ISL_742966\|2020 | ···················· | ···················· | ···················· | ···················· | ···················· | ···················· |
| Z | 3466 hCoV-19/Wales/PHWC-49E1D3/2020\|EPI_ISL_742967\|2020 | ···················· | ···················· | ···················· | ···················· | ···················· | ···················· |
| Z | 3467 hCoV-19/Wales/PHWC-49E1E2/2020\|EPI_ISL_742968\|2020 | ···················· | ···················· | ···················· | ···················· | ···················· | ···················· |
| Z | 3468 hCoV-19/Wales/PHWC-49E20D/2020\|EPI_ISL_742969\|2020 | ···················· | ···················· | ···················· | ···················· | ···················· | ···················· |
| Z | 3469 hCoV-19/Wales/PHWC-49E23A/2020\|EPI_ISL_742972\|2020 | ···················· | ···················· | ···················· | ···················· | ···················· | ···················· |
| Z | 3470 hCoV-19/Wales/PHWC-49E249/2020\|EPI_ISL_742973\|2020 | ···················· | ···················· | ···················· | ···················· | ···················· | ···················· |
| Z | 3471 hCoV-19/Wales/PHWC-49E258/2020\|EPI_ISL_742974\|2020 | ···················· | ···················· | ···················· | ···················· | ···················· | ···················· |
| Z | 3472 hCoV-19/Wales/PHWC-49E267/2020\|EPI_ISL_742975\|2020 | ···················· | ···················· | ···················· | ···················· | ···················· | ···················· |
| Z | 3473 hCoV-19/Wales/PHWC-49E285/2020\|EPI_ISL_742977\|2020 | ···················· | ···················· | ···················· | ···················· | ···················· | ···················· |
| Z | 3474 hCoV-19/Wales/PHWC-49E294/2020\|EPI_ISL_742978\|2020 | ···················· | ···················· | ···················· | ···················· | ···················· | ···················· |
| Z | 3475 hCoV-19/Wales/PHWC-49E2A3/2020\|EPI_ISL_742979\|2020 | ···················· | ···················· | ···················· | ···················· | ···················· | ···················· |
| Z | 3476 hCoV-19/India/AP-CCMB-BB648/2020\|EPI_ISL_745201\|20 | ···················· | ·A·················· | ···················· | ···················· | ···················· | ···A················ |
| Z | 3477 hCoV-19/Belgium/UGent-313/2020\|EPI_ISL_745202\|2020 | ···················· | ···················· | ···················· | ···················· | ···················· | ···················· |
| Z | 3478 hCoV-19/Belgium/UGent-314/2020\|EPI_ISL_745203\|2020 | ···················· | ···················· | ···················· | ···················· | ···················· | ···················· |
| Z | 3480 hCoV-19/Belgium/UGent-317/2020\|EPI_ISL_745206\|2020 | ···················· | ···················· | ···················· | ···················· | ···················· | ···················· |
| Z | 3481 hCoV-19/Belgium/UGent-318/2020\|EPI_ISL_745207\|2020 | ···················· | ···················· | ···················· | ···················· | ···················· | ···················· |
| Z | 3485 hCoV-19/Belgium/UGent-327/2020\|EPI_ISL_745214\|2020 | ···················· | ···················· | ···················· | ···················· | ···················· | ···················· |
| Z | 3486 hCoV-19/Belgium/UGent-328/2020\|EPI_ISL_745215\|2020 | ···················· | ···················· | ···················· | ···················· | ···················· | ···················· |
| Z | 3487 hCoV-19/Belgium/UGent-330/2020\|EPI_ISL_745216\|2020 | ···················· | ···················· | ···················· | ···················· | ···················· | ···················· |
| Z | 3488 hCoV-19/Belgium/UGent-332/2020\|EPI_ISL_745217\|2020 | ···················· | ···················· | ···················· | ···················· | ···················· | ···················· |
| Z | 3491 hCoV-19/Belgium/rega-12141317/2020\|EPI_ISL_745222\| | ···················· | ···················· | ···················· | ···················· | ···················· | ···················· |
| Z | 3492 hCoV-19/Russia/MOS-CRIE-7182855/2020\|EPI_ISL_74522 | ···················· | ···················· | ···················· | ···················· | ···················· | ···················· |
| Z | 3493 hCoV-19/Russia/MOS-CRIE-13476745/2020\|EPI_ISL_7452 | ···················· | ···················· | ···················· | ···················· | ···················· | ···················· |
| Z | 3494 hCoV-19/Russia/MOS-CRIE-13370222/2020\|EPI_ISL_7452 | ···················· | ···················· | ···················· | ···················· | ···················· | ···················· |
| Z | 3495 hCoV-19/Russia/MOS-CRIE-13384497/2020\|EPI_ISL_7452 | ···················· | ···················· | ···················· | ···················· | ···················· | ···················· |
| Z | 3496 hCoV-19/Russia/MOS-CRIE-13372214/2020\|EPI_ISL_7452 | ················G··· | ···················· | ···················· | ···················· | ···················· | ···················· |
| Z | 3497 hCoV-19/Russia/MOS-CRIE-13386480/2020\|EPI_ISL_7452 | ···················· | ···················· | ···················· | ···················· | ···················· | ···················· |
| Z | 3498 hCoV-19/Russia/MOS-CRIE-13373431/2020\|EPI_ISL_7452 | ···················· | ···················· | ···················· | ···················· | ···················· | ···················· |
| Z | 3499 hCoV-19/Russia/MOS-CRIE-13381035/2020\|EPI_ISL_7452 | ···················· | ···················· | ···················· | ···················· | ···················· | ···················· |
| Z | 3500 hCoV-19/Russia/MOS-CRIE-13380178/2020\|EPI_ISL_7452 | ···················· | ···················· | ···················· | ···················· | ···················· | ···················· |
| Z | 3501 hCoV-19/USA/TX-DSHS-1830/2020\|EPI_ISL_745276\|2020- | ···················· | ···················· | ···················· | ···················· | ···················· | ···················· |
| Z | 3502 hCoV-19/USA/TX-DSHS-1831/2020\|EPI_ISL_745277\|2020- | ···················· | ···················· | ···················· | ···················· | ···················· | ···················· |
| Z | 3503 hCoV-19/USA/TX-DSHS-1832/2020\|EPI_ISL_745278\|2020- | ···················· | ···················· | ···················· | ···················· | ···················· | ···················· |
| Z | 3504 hCoV-19/USA/TX-DSHS-1833/2020\|EPI_ISL_745279\|2020- | ···················· | ···················· | ···················· | ···················· | ···················· | ···················· |
| Z | 3505 hCoV-19/Chile/AR-178998/2020\|EPI_ISL_746500\|2020-0 | ···················· | ···················· | ···················· | ···················· | ···················· | ···················· |
| Z | 3506 hCoV-19/Chile/AR-192896/2020\|EPI_ISL_746501\|2020-0 | ···················· | ···················· | ···················· | ···················· | ···················· | ···················· |
| Z | 3507 hCoV-19/Chile/AR-192900/2020\|EPI_ISL_746502\|2020-0 | ···················· | ···················· | ···················· | ···················· | ···················· | ···················· |
| Z | 3508 hCoV-19/Chile/AR-208475/2020\|EPI_ISL_746503\|2020-1 | ···················· | ···················· | ···················· | ···················· | ···················· | ···················· |
| Z | 3509 hCoV-19/Chile/AR-208476/2020\|EPI_ISL_746504\|2020-1 | ···················· | ···················· | ···················· | ···················· | ···················· | ···················· |
| Z | 3510 hCoV-19/Chile/AR-208477/2020\|EPI_ISL_746505\|2020-1 | ···················· | ···················· | ···················· | ···················· | ···················· | ···················· |
| Z | 3511 hCoV-19/Chile/BI-109143/2020\|EPI_ISL_746506\|2020-0 | ···················· | ···················· | ···················· | ···················· | ···················· | ···················· |
| Z | 3512 hCoV-19/Chile/BI-167039/2020\|EPI_ISL_746507\|2020-0 | ···················· | ···················· | ···················· | ···················· | ···················· | ···················· |
| Z | 3513 hCoV-19/Chile/BI-167040/2020\|EPI_ISL_746508\|2020-0 | ···················· | ···················· | ···················· | ···················· | ···················· | ···················· |
| Z | 3514 hCoV-19/Chile/BI-178219/2020\|EPI_ISL_746509\|2020-0 | ···················· | ···················· | ···················· | ···················· | ···················· | ···················· |
| Z | 3515 hCoV-19/Chile/BI-185934/2020\|EPI_ISL_746510\|2020-0 | ···················· | ···················· | ···················· | ···················· | ···················· | ···················· |
| Z | 3516 hCoV-19/Chile/BI-194033/2020\|EPI_ISL_746511\|2020-0 | ···················· | ···················· | ···················· | ···················· | ···················· | ···················· |

| | | | | | | | |
|---|---|---|---|---|---|---|---|
| Z | 3517 hCoV-19/Chile/BI-194034/2020\|EPI_ISL_746512\|2020-0 | ···················· | ···················· | ····················· | ···················· | ···················· | ···················· |
| Z | 3518 hCoV-19/Chile/BI-199845/2020\|EPI_ISL_746513\|2020-1 | ···················· | ···················· | ····················· | ···················· | ···················· | ···················· |
| Z | 3519 hCoV-19/Chile/BI-199848/2020\|EPI_ISL_746514\|2020-1 | ···················· | ···················· | ····················· | ···················· | ···················· | ···················· |
| Z | 3520 hCoV-19/Chile/BI-199849/2020\|EPI_ISL_746515\|2020-1 | ···················· | ···················· | ····················· | ···················· | ···················· | ···················· |
| Z | 3521 hCoV-19/Chile/BI-199851/2020\|EPI_ISL_746516\|2020-1 | ···················· | ···················· | ····················· | ···················· | ···················· | ···················· |
| Z | 3522 hCoV-19/Chile/BI-206407/2020\|EPI_ISL_746517\|2020-1 | ···················· | ···················· | ····················· | ···················· | ···················· | ···················· |
| Z | 3523 hCoV-19/Chile/BI-206419/2020\|EPI_ISL_746518\|2020-1 | ···················· | ···················· | ···T················· | ···················· | ···················· | ···················· |
| Z | 3524 hCoV-19/Chile/BI-206420/2020\|EPI_ISL_746519\|2020-1 | ···················· | ···················· | ····················· | ···················· | ···················· | ···················· |
| Z | 3525 hCoV-19/Chile/MA-164318/2020\|EPI_ISL_746520\|2020-0 | ···················· | ···················· | ····················· | ···················· | ···················· | ···················· |
| Z | 3526 hCoV-19/Chile/MA-194107-B/2020\|EPI_ISL_746521\|2020 | ···················· | ···················· | ····················· | ···················· | ···················· | ···················· |
| Z | 3527 hCoV-19/Chile/MA-194108-B/2020\|EPI_ISL_746522\|2020 | ···················· | ···················· | ····················· | ···················· | ···················· | ···················· |
| Z | 3528 hCoV-19/Chile/MA-194109-B/2020\|EPI_ISL_746523\|2020 | ···················· | ···················· | ····················· | ···················· | ···················· | ···················· |
| Z | 3529 hCoV-19/Chile/MA-194111-B/2020\|EPI_ISL_746524\|2020 | ···················· | ···················· | ····················· | ···················· | ···················· | ···················· |
| Z | 3530 hCoV-19/Chile/MA-194112-B/2020\|EPI_ISL_746525\|2020 | ···················· | ···················· | ····················· | ···················· | ···················· | ···················· |
| Z | 3531 hCoV-19/Chile/MA-194113-B/2020\|EPI_ISL_746526\|2020 | ···················· | ···················· | ····················· | ···················· | ···················· | ···················· |
| Z | 3532 hCoV-19/Chile/MA-194114-B/2020\|EPI_ISL_746527\|2020 | ···················· | ···················· | ····················· | ···················· | ···················· | ···················· |
| Z | 3533 hCoV-19/Chile/MA-194115-B/2020\|EPI_ISL_746528\|2020 | ···················· | ···················· | ····················· | ···················· | ···················· | ···················· |
| Z | 3534 hCoV-19/Chile/MA-194116-B/2020\|EPI_ISL_746529\|2020 | ···················· | ···················· | ····················· | ···················· | ···················· | ···················· |
| Z | 3535 hCoV-19/Chile/MA-201640/2020\|EPI_ISL_746530\|2020-1 | ···················· | ···················· | ····················· | ···················· | ···················· | ···················· |
| Z | 3536 hCoV-19/Chile/MA-201641/2020\|EPI_ISL_746531\|2020-1 | ···················· | ···················· | ····················· | ···················· | ···················· | ···················· |
| Z | 3537 hCoV-19/Chile/MA-212210/2020\|EPI_ISL_746532\|2020-1 | ···················· | ···················· | ····················· | ···················· | ···················· | ···················· |
| Z | 3538 hCoV-19/Chile/MA-212211/2020\|EPI_ISL_746533\|2020-1 | ············T······· | ·········A·········· | ····················· | ···················· | ···················· | ···················· |
| Z | 3539 hCoV-19/Chile/MA-212212/2020\|EPI_ISL_746534\|2020-1 | ···················· | ···················· | ····················· | ···················· | ···················· | ···················· |
| Z | 3540 hCoV-19/Chile/MA-212213/2020\|EPI_ISL_746535\|2020-1 | ···················· | ···················· | ····················· | ···················· | ···················· | ···················· |
| Z | 3541 hCoV-19/Chile/NB-175519/2020\|EPI_ISL_746536\|2020-0 | ···················· | ···················· | ····················· | ···················· | ···················· | ···················· |
| Z | 3542 hCoV-19/Chile/NB-175522/2020\|EPI_ISL_746537\|2020-0 | ···················· | ···················· | ····················· | ···················· | ···················· | ···················· |
| Z | 3543 hCoV-19/Chile/NB-196351/2020\|EPI_ISL_746538\|2020-1 | ···················· | ···················· | ····················· | ···················· | ···················· | ···················· |
| Z | 3544 hCoV-19/Chile/NB-196353/2020\|EPI_ISL_746539\|2020-1 | ···················· | ···················· | ····················· | ···················· | ···················· | ···················· |
| Z | 3545 hCoV-19/Chile/NB-202522/2020\|EPI_ISL_746540\|2020-1 | ···················· | ···················· | ····················· | ···················· | ···················· | ···················· |
| Z | 3546 hCoV-19/Chile/NB-202524/2020\|EPI_ISL_746541\|2020-1 | ···················· | ···················· | ····················· | ···················· | ···················· | ···················· |
| Z | 3547 hCoV-19/Chile/NB-202530/2020\|EPI_ISL_746542\|2020-1 | ···················· | ···················· | ····················· | ···················· | ···················· | ···················· |
| Z | 3548 hCoV-19/Chile/NB-207884/2020\|EPI_ISL_746543\|2020-1 | ···················· | ···················· | ····················· | ···················· | ···················· | ···················· |
| Z | 3549 hCoV-19/Chile/NB-207885/2020\|EPI_ISL_746544\|2020-1 | ···················· | ···················· | ····················· | ···················· | ···················· | ···················· |
| Z | 3550 hCoV-19/Chile/RM-108589/2020\|EPI_ISL_746545\|2020-0 | ···················· | ···················· | ····················· | ···················· | ···················· | ···················· |
| Z | 3551 hCoV-19/Chile/RM-108734/2020\|EPI_ISL_746546\|2020-0 | ···················· | ···················· | ····················· | ···················· | ···················· | ···················· |
| Z | 3552 hCoV-19/Chile/RM-108736/2020\|EPI_ISL_746547\|2020-0 | ···················· | ···················· | ····················· | ···················· | ···················· | ···················· |
| Z | 3553 hCoV-19/Chile/RM-108996/2020\|EPI_ISL_746548\|2020-0 | ···················· | ···················· | ····················· | ···················· | ···················· | ···················· |
| Z | 3554 hCoV-19/Chile/RM-108997/2020\|EPI_ISL_746549\|2020-0 | ···················· | ···················· | ····················· | ···················· | ···················· | ···················· |
| Z | 3555 hCoV-19/Chile/RM-108999/2020\|EPI_ISL_746550\|2020-0 | ···················· | ···················· | ····················· | ···················· | ···················· | ···················· |
| Z | 3556 hCoV-19/Chile/RM-109139/2020\|EPI_ISL_746551\|2020-0 | ···················· | ···················· | ····················· | ···················· | ···················· | ···················· |
| Z | 3557 hCoV-19/Chile/RM-109226/2020\|EPI_ISL_746552\|2020-0 | ···················· | ···················· | ····················· | ···················· | ···················· | ···················· |
| Z | 3558 hCoV-19/Chile/RM-109231/2020\|EPI_ISL_746553\|2020-0 | ···················· | ···················· | ····················· | ···················· | ···················· | ···················· |
| Z | 3559 hCoV-19/Chile/RM-109455/2020\|EPI_ISL_746554\|2020-0 | ···················· | ···················· | ····················· | ···················· | ···················· | ···················· |
| Z | 3560 hCoV-19/Chile/RM-109521/2020\|EPI_ISL_746555\|2020-0 | ···················· | ···················· | ····················· | ···················· | ···················· | ···················· |
| Z | 3561 hCoV-19/Chile/RM-109567/2020\|EPI_ISL_746556\|2020-0 | ···················· | ···················· | ····················· | ···················· | ···················· | ···················· |
| Z | 3562 hCoV-19/Chile/RM-109578/2020\|EPI_ISL_746557\|2020-0 | ···················· | ···················· | ····················· | ···················· | ···················· | ···················· |
| Z | 3563 hCoV-19/Chile/RM-109586/2020\|EPI_ISL_746558\|2020-0 | ···················· | ···················· | ····················· | ···················· | ···················· | ···················· |
| Z | 3564 hCoV-19/Chile/RM-109587/2020\|EPI_ISL_746559\|2020-0 | ···················· | ···················· | ····················· | ···················· | ···················· | ···················· |
| Z | 3565 hCoV-19/Chile/RM-109600/2020\|EPI_ISL_746560\|2020-0 | ···················· | ···················· | ····················· | ···················· | ···················· | ···················· |
| Z | 3566 hCoV-19/Chile/RM-109602/2020\|EPI_ISL_746561\|2020-0 | ···················· | ···················· | ····················· | ···················· | ···················· | ···················· |
| Z | 3567 hCoV-19/Chile/RM-110507/2020\|EPI_ISL_746562\|2020-0 | ···················· | ···················· | ····················· | ···················· | ···················· | ···················· |
| Z | 3568 hCoV-19/Chile/RM-110508/2020\|EPI_ISL_746563\|2020-0 | ···················· | ···················· | ····················· | ···················· | ···················· | ···················· |
| Z | 3569 hCoV-19/Chile/RM-110525/2020\|EPI_ISL_746564\|2020-0 | ···················· | ···················· | ····················· | ···················· | ···················· | ···················· |
| Z | 3570 hCoV-19/Chile/RM-110750/2020\|EPI_ISL_746565\|2020-0 | ···················· | ···················· | ····················· | ···················· | ···················· | ···················· |
| Z | 3571 hCoV-19/Chile/RM-110802/2020\|EPI_ISL_746566\|2020-0 | ···················· | ···················· | ····················· | ···················· | ···················· | ···················· |
| Z | 3572 hCoV-19/Chile/RM-110804/2020\|EPI_ISL_746567\|2020-0 | ···················· | ···················· | ····················· | ···················· | ···················· | ···················· |
| Z | 3573 hCoV-19/Chile/RM-110815/2020\|EPI_ISL_746568\|2020-0 | ···················· | ···················· | ····················· | ···················· | ···················· | ···················· |
| Z | 3574 hCoV-19/Chile/RM-110874/2020\|EPI_ISL_746569\|2020-0 | ···················· | ···················· | ····················· | ···················· | ···················· | ···················· |
| Z | 3575 hCoV-19/Chile/RM-112763/2020\|EPI_ISL_746590\|2020-0 | ···················· | ···················· | ····················· | ···················· | ···················· | ···················· |
| Z | 3576 hCoV-19/Chile/RM-112769/2020\|EPI_ISL_746591\|2020-0 | ···················· | ···················· | ····················· | ···················· | ···················· | ···················· |
| Z | 3577 hCoV-19/Chile/RM-112773/2020\|EPI_ISL_746592\|2020-0 | ···················· | ···················· | ····················· | ···················· | ···················· | ···················· |
| Z | 3578 hCoV-19/Chile/RM-112777/2020\|EPI_ISL_746593\|2020-0 | ···················· | ···················· | ····················· | ···················· | ···················· | ···················· |
| Z | 3579 hCoV-19/Chile/RM-126528/2020\|EPI_ISL_746594\|2020-0 | ···················· | ···················· | ····················· | ···················· | ···················· | ···················· |
| Z | 3580 hCoV-19/Chile/RM-126534/2020\|EPI_ISL_746595\|2020-0 | ···················· | ···················· | ····················· | ···················· | ···················· | ···················· |
| Z | 3581 hCoV-19/Chile/RM-126787/2020\|EPI_ISL_746596\|2020-0 | ···················· | ···················· | ····················· | ···················· | ···················· | ···················· |
| Z | 3582 hCoV-19/Chile/RM-127204/2020\|EPI_ISL_746597\|2020-0 | ···················· | ···················· | ····················· | ···················· | ···················· | ···················· |
| Z | 3583 hCoV-19/Chile/RM-127405/2020\|EPI_ISL_746598\|2020-0 | ···················· | ···················· | ····················· | ···················· | ···················· | ···················· |
| Z | 3584 hCoV-19/Chile/RM-127424/2020\|EPI_ISL_746599\|2020-0 | ···················· | ···················· | ····················· | ···················· | ···················· | ···················· |
| Z | 3585 hCoV-19/USA/CA-CZB-14580/2020\|EPI_ISL_739400\|2020- | ···················· | ···················· | ····················· | ···················· | ···················· | ···················· |
| Z | 3586 hCoV-19/USA/CA-CZB-15442/2020\|EPI_ISL_739401\|2020- | ···················· | ···················· | ····················· | ···················· | ···················· | ···················· |
| Z | 3587 hCoV-19/USA/CA-CZB-15333/2020\|EPI_ISL_739402\|2020- | ···················· | ···················· | ····················· | ···················· | ···················· | ···················· |
| Z | 3588 hCoV-19/USA/CA-CZB-14827/2020\|EPI_ISL_739403\|2020- | ···················· | ···················· | ····················· | ···················· | ···················· | ···················· |
| Z | 3589 hCoV-19/USA/CA-CZB-15055/2020\|EPI_ISL_739404\|2020- | ···················· | ·A·················· | ····················· | ···················· | ···················· | ···················· |
| Z | 3590 hCoV-19/USA/CA-CZB-14826/2020\|EPI_ISL_739405\|2020- | ···················· | ···················· | ····················· | ···················· | ···················· | ···················· |
| Z | 3591 hCoV-19/USA/CA-CZB-13802/2020\|EPI_ISL_739406\|2020- | ···················· | ·A·················· | ····················· | ···················· | ···················· | ···················· |
| Z | 3593 hCoV-19/USA/CA-CZB-13974/2020\|EPI_ISL_739408\|2020- | ···················· | ···················· | ····················· | ···················· | ···················· | ···················· |
| Z | 3594 hCoV-19/USA/CA-CZB-15002/2020\|EPI_ISL_739409\|2020- | ···················· | ···················· | ····················· | ···················· | ···················· | ···················· |
| Z | 3595 hCoV-19/USA/CA-CZB-14669/2020\|EPI_ISL_739410\|2020- | ···················· | ···················· | ····················· | ···················· | ···················· | ···················· |
| Z | 3596 hCoV-19/USA/CA-CZB-14772/2020\|EPI_ISL_739411\|2020- | ···················· | ···················· | ····················· | ···················· | ···················· | ···················· |
| Z | 3597 hCoV-19/USA/CA-CZB-14676/2020\|EPI_ISL_739412\|2020- | ···················· | ···················· | ····················· | ···················· | ···················· | ···················· |

| | | | | | | | | | | |
|---|---|---|---|---|---|---|---|---|---|---|
| Z | 3598 | hCoV-19/USA/CA-CZB-15100/2020\|EPI_ISL_739413\|2020- | ···················· | ···················· | ···················· | ···················· | ···················· | ···················· | | |
| Z | 3599 | hCoV-19/USA/CA-CZB-15362/2020\|EPI_ISL_739414\|2020- | ···················· | ···················· | ···················· | ···················· | ···················· | ···················· | | |
| Z | 3600 | hCoV-19/USA/CA-CZB-15254/2020\|EPI_ISL_739415\|2020- | ···················· | ···················· | ···················· | ···················· | ···················· | ···················· | | |
| Z | 3601 | hCoV-19/USA/CA-CZB-15156/2020\|EPI_ISL_739416\|2020- | ···················· | ·A·················· | ···················· | ···················· | ···················· | ···················· | | |
| Z | 3602 | hCoV-19/USA/CA-CZB-14831/2020\|EPI_ISL_739417\|2020- | ···················· | ···················· | ···················· | ···················· | ···················· | ···················· | | |
| Z | 3603 | hCoV-19/USA/CA-CZB-14852/2020\|EPI_ISL_739418\|2020- | ···················· | ···················· | ···················· | ···················· | ···················· | ···················· | | |
| Z | 3604 | hCoV-19/USA/CA-CZB-15975/2020\|EPI_ISL_739419\|2020- | ···················· | ·A·················· | ···················· | ···················· | ···················· | ···················· | | |
| Z | 3605 | hCoV-19/USA/CA-CZB-13730/2020\|EPI_ISL_739420\|2020- | ···················· | ···················· | ···················· | ···················· | ···················· | ···················· | | |
| Z | 3606 | hCoV-19/USA/CA-CZB-15470/2020\|EPI_ISL_739421\|2020- | ···················· | ···················· | ···T················ | ···················· | ···················· | ···················· | | |
| Z | 3607 | hCoV-19/USA/CA-CZB-14351/2020\|EPI_ISL_739422\|2020- | ···················· | ····A··············· | ···················· | ···················· | ···················· | ···················· | | |
| Z | 3608 | hCoV-19/USA/CA-CZB-13723/2020\|EPI_ISL_739423\|2020- | ···················· | ···················· | ···················· | ···················· | ···················· | ···················· | | |
| Z | 3610 | hCoV-19/USA/CA-CZB-14838/2020\|EPI_ISL_739425\|2020- | ···················· | ···················· | ···················· | ···················· | ···················· | ···················· | | |
| Z | 3612 | hCoV-19/USA/CA-CZB-14243/2020\|EPI_ISL_739427\|2020- | ···················· | ·A·················· | ···················· | ···················· | ···················· | ···················· | | |
| Z | 3613 | hCoV-19/USA/CA-CZB-14693/2020\|EPI_ISL_739428\|2020- | ···················· | ···················· | ···················· | ···················· | ···················· | ···················· | | |
| Z | 3614 | hCoV-19/USA/CA-CZB-14158/2020\|EPI_ISL_739429\|2020- | ···················· | ·A·················· | ···················· | ···················· | ···················· | ···················· | | |
| Z | 3615 | hCoV-19/USA/CA-CZB-14753/2020\|EPI_ISL_739430\|2020- | ···················· | ···················· | ···················· | ···················· | ···················· | ···················· | | |
| Z | 3617 | hCoV-19/USA/CA-CZB-15460/2020\|EPI_ISL_739432\|2020- | ···················· | ···················· | ···················· | ···················· | ···················· | ···················· | | |
| Z | 3618 | hCoV-19/USA/CA-CZB-14755/2020\|EPI_ISL_739433\|2020- | ···················· | ···················· | ···················· | ···················· | ···················· | ···················· | | |
| Z | 3619 | hCoV-19/USA/CA-CZB-14410/2020\|EPI_ISL_739434\|2020- | ···················· | ···················· | ···················· | ···················· | ···················· | ···················· | | |
| Z | 3620 | hCoV-19/USA/CA-CZB-13978/2020\|EPI_ISL_739435\|2020- | ···················· | ···················· | ···················· | ···················· | ···················· | ···················· | | |
| Z | 3621 | hCoV-19/USA/CA-CZB-14806/2020\|EPI_ISL_739436\|2020- | ···················· | ···················· | ···················· | ···················· | ···················· | ···················· | | |
| Z | 3622 | hCoV-19/USA/CA-CZB-14945/2020\|EPI_ISL_739437\|2020- | ···················· | ···················· | ···················· | ···················· | ···················· | ···················· | | |
| Z | 3623 | hCoV-19/USA/CA-CZB-14646/2020\|EPI_ISL_739438\|2020- | ···················· | ···················· | ···················· | ···················· | ···················· | ···················· | | |
| Z | 3624 | hCoV-19/USA/CA-CZB-13987/2020\|EPI_ISL_739439\|2020- | ···················· | ·A·················· | ···················· | ···················· | ···················· | ···················· | | |
| Z | 3625 | hCoV-19/USA/CA-CZB-14869/2020\|EPI_ISL_739445\|2020- | ···················· | ···················· | ···················· | ···················· | ···················· | ···················· | | |
| Z | 3626 | hCoV-19/USA/CA-CZB-13984/2020\|EPI_ISL_739446\|2020- | ···················· | ···················· | ···················· | ···················· | ···················· | ···················· | | |
| Z | 3627 | hCoV-19/USA/CA-CZB-14793/2020\|EPI_ISL_739447\|2020- | ···················· | ···················· | ···················· | ···················· | ···················· | ···················· | | |
| Z | 3630 | hCoV-19/USA/CA-CZB-14907/2020\|EPI_ISL_739450\|2020- | ···················· | ·A·················· | ···················· | ···················· | ···················· | ···················· | | |
| Z | 3631 | hCoV-19/USA/CA-CZB-14512/2020\|EPI_ISL_739451\|2020- | ···················· | ·A·················· | ···················· | ···················· | ···················· | ···················· | | |
| Z | 3632 | hCoV-19/USA/CA-CZB-14851/2020\|EPI_ISL_739452\|2020- | ···················· | ···················· | ···················· | ···················· | ···················· | ···················· | | |
| Z | 3634 | hCoV-19/USA/CA-CZB-13952/2020\|EPI_ISL_739454\|2020- | ···················· | ···················· | ···················· | ···················· | ···················· | ···················· | | |
| Z | 3635 | hCoV-19/USA/CA-CZB-14928/2020\|EPI_ISL_739455\|2020- | ···················· | ·A·················· | ···················· | ···················· | ···················· | ···················· | | |
| Z | 3636 | hCoV-19/USA/CA-CZB-15238/2020\|EPI_ISL_739456\|2020- | ···················· | ···················· | ···················· | ···················· | ···················· | ···················· | | |
| Z | 3637 | hCoV-19/USA/CA-CZB-15320/2020\|EPI_ISL_739457\|2020- | ···················· | ·A·················· | ···················· | ···················· | ···················· | ···················· | | |
| Z | 3638 | hCoV-19/USA/CA-CZB-15159/2020\|EPI_ISL_739458\|2020- | ···················· | ···················· | ···················· | ···················· | ···················· | ···················· | | |
| Z | 3639 | hCoV-19/USA/CA-CZB-14390/2020\|EPI_ISL_739459\|2020- | ···················· | ···················· | ···················· | ···················· | N·················NN | ···················· | 1 | 1 |
| Z | 3640 | hCoV-19/USA/CA-CZB-15476/2020\|EPI_ISL_739460\|2020- | ···················· | ···················· | ···T················ | ···················· | ···················· | ···················· | | |
| Z | 3641 | hCoV-19/USA/CA-CZB-14837/2020\|EPI_ISL_739461\|2020- | ···················· | ···················· | ···················· | ···················· | ···················· | ···················· | | |
| Z | 3642 | hCoV-19/USA/CA-CZB-15432/2020\|EPI_ISL_739462\|2020- | ···················· | ···················· | ···················· | ···················· | ···················· | ···················· | | |
| Z | 3643 | hCoV-19/USA/CA-CZB-14409/2020\|EPI_ISL_739463\|2020- | ···················· | ···················· | ···················· | ···················· | ···················· | ···················· | | |
| Z | 3644 | hCoV-19/USA/CA-CZB-15401/2020\|EPI_ISL_739464\|2020- | ···················· | ···················· | ···················· | ···················· | ···················· | ···················· | | |
| Z | 3645 | hCoV-19/USA/CA-CZB-14667/2020\|EPI_ISL_739465\|2020- | ···················· | ···················· | ···················· | ···················· | NNN················· | ···················· | | 1 |
| Z | 3646 | hCoV-19/USA/CA-CZB-16075/2020\|EPI_ISL_739466\|2020- | ···················· | ·A·················· | ···················· | ···················· | ···················· | ···················· | | |
| Z | 3647 | hCoV-19/USA/CA-CZB-16154/2020\|EPI_ISL_739467\|2020- | ···················· | ·A·················· | ···················· | ···················· | ···················· | ···················· | | |
| Z | 3648 | hCoV-19/USA/CA-CZB-14114/2020\|EPI_ISL_739468\|2020- | ···················· | ···················· | ···················· | ···················· | ···················· | ···················· | | |
| Z | 3649 | hCoV-19/USA/CA-CZB-11271/2020\|EPI_ISL_739469\|2020- | ···················· | ···················· | ···················· | ···················· | ···················· | ···················· | | |
| Z | 3650 | hCoV-19/USA/CA-CZB-13865/2020\|EPI_ISL_739470\|2020- | ···················· | ···················· | ···················· | ···················· | ···················· | ···················· | | |
| Z | 3651 | hCoV-19/USA/CA-CZB-11262/2020\|EPI_ISL_739471\|2020- | ···················· | ···················· | ···················· | ···················· | ···················· | ···················· | | |
| Z | 3652 | hCoV-19/USA/CA-CZB-13867/2020\|EPI_ISL_739472\|2020- | ···················· | ·A·················· | ···················· | ···················· | ···················· | ···················· | | |
| Z | 3653 | hCoV-19/USA/CA-CZB-14254/2020\|EPI_ISL_739473\|2020- | ···················· | ···················· | ···················· | ···················· | ···················· | ···················· | | |
| Z | 3654 | hCoV-19/USA/CA-CZB-14605/2020\|EPI_ISL_739474\|2020- | ···················· | ···················· | ···················· | ···················· | ···················· | ···················· | | |
| Z | 3655 | hCoV-19/USA/CA-CZB-14830/2020\|EPI_ISL_739475\|2020- | ···················· | ···················· | ···················· | ···················· | ···················· | ···················· | | |
| Z | 3656 | hCoV-19/USA/CA-CZB-14568/2020\|EPI_ISL_739476\|2020- | ···················· | ·A·················· | ···················· | ···················· | ···················· | ···················· | | |
| Z | 3657 | hCoV-19/USA/CA-CZB-13728/2020\|EPI_ISL_739477\|2020- | ···················· | ···················· | ···················· | ···················· | ···················· | ···················· | | |
| Z | 3658 | hCoV-19/USA/CA-CZB-16175/2020\|EPI_ISL_739478\|2020- | ···················· | ···················· | ···················· | ···················· | ···················· | ···················· | | |
| Z | 3659 | hCoV-19/USA/CA-CZB-15057/2020\|EPI_ISL_739479\|2020- | ···················· | ·A·················· | ···················· | ···················· | ···················· | ···················· | | |
| Z | 3660 | hCoV-19/Luxembourg/LNS3993810/2020\|EPI_ISL_740400\| | ···················· | ···················· | ···················· | ···················· | ···················· | ···················· | | |
| Z | 3661 | hCoV-19/Luxembourg/LNS4007359/2020\|EPI_ISL_740401\| | ···················· | ···················· | ···················· | ···················· | ···················· | ···················· | | |
| Z | 3662 | hCoV-19/Luxembourg/LNS4009709/2020\|EPI_ISL_740402\| | ···················· | ···················· | ···················· | ···················· | ···················· | ···················· | | |
| Z | 3663 | hCoV-19/Luxembourg/LNS4009932/2020\|EPI_ISL_740403\| | ···················· | ···················· | ···················· | ···················· | ···················· | ···················· | | |
| Z | 3664 | hCoV-19/Luxembourg/LNS4028717/2020\|EPI_ISL_740404\| | ···················· | ···················· | ···················· | ···················· | ···················· | ···················· | | |
| Z | 3665 | hCoV-19/Luxembourg/LNS4040746/2020\|EPI_ISL_740405\| | ···················· | ···················· | ···················· | ···················· | ···················· | ···················· | | |
| Z | 3666 | hCoV-19/Luxembourg/LNS4041175/2020\|EPI_ISL_740406\| | ···················· | ···················· | ···················· | ···················· | ···················· | ···················· | | |
| Z | 3667 | hCoV-19/Luxembourg/LNS4044086/2020\|EPI_ISL_740407\| | ···················· | ···················· | ···················· | ···················· | ···················· | ···················· | | |
| Z | 3668 | hCoV-19/Luxembourg/LNS4050385/2020\|EPI_ISL_740410\| | ···················· | ···················· | ···················· | ···················· | ···················· | ···················· | | |
| Z | 3669 | hCoV-19/Luxembourg/LNS4050425/2020\|EPI_ISL_740411\| | ···················· | G··················· | ···················· | ···················· | ···················· | ···················· | | |
| Z | 3670 | hCoV-19/Luxembourg/LNS4055666/2020\|EPI_ISL_740412\| | ···················· | ···················· | ···················· | ···················· | ···················· | ···················· | | |
| Z | 3671 | hCoV-19/Luxembourg/LNS4062347/2020\|EPI_ISL_740413\| | ···················· | ···················· | ···················· | ···················· | ···················· | ···················· | | |
| Z | 3672 | hCoV-19/Luxembourg/LNS4063851/2020\|EPI_ISL_740414\| | ···················· | ···················· | ···················· | ···················· | ···················· | ···················· | | |
| Z | 3673 | hCoV-19/Luxembourg/LNS4071178/2020\|EPI_ISL_740415\| | ···················· | ···················· | ···················· | ···················· | ···················· | ···················· | | |
| Z | 3674 | hCoV-19/Luxembourg/LNS4075909/2020\|EPI_ISL_740416\| | ···················· | ···················· | ···················· | ···················· | ···················· | ···················· | | |
| Z | 3675 | hCoV-19/Luxembourg/LNS4079524/2020\|EPI_ISL_740417\| | ···················· | ···················· | ···················· | ···················· | ···················· | ···················· | | |
| Z | 3676 | hCoV-19/Luxembourg/LNS4081363/2020\|EPI_ISL_740418\| | ···················· | ···················· | ···················· | ···················· | ···················· | ···················· | | |
| Z | 3677 | hCoV-19/Luxembourg/LNS4081687/2020\|EPI_ISL_740419\| | ···················· | ···················· | ···················· | ···················· | ···················· | ···················· | | |
| Z | 3678 | hCoV-19/Luxembourg/LNS4085967/2020\|EPI_ISL_740420\| | ···················· | ···················· | ···················· | ···················· | ···················· | ···················· | | |
| Z | 3679 | hCoV-19/Luxembourg/LNS4101182/2020\|EPI_ISL_740422\| | ···················· | ···················· | ···················· | ···················· | ···················· | ···················· | | |
| Z | 3680 | hCoV-19/Luxembourg/LNS4109159/2020\|EPI_ISL_740423\| | ···················· | ···················· | ···················· | ···················· | ···················· | ···················· | | |
| Z | 3681 | hCoV-19/Luxembourg/LNS4109804/2020\|EPI_ISL_740424\| | ···················· | ···················· | ···················· | ···················· | ···················· | ···················· | | |
| Z | 3682 | hCoV-19/Luxembourg/LNS4136398/2020\|EPI_ISL_740425\| | ···················· | ···················· | ···················· | ···················· | ···················· | ···················· | | |
| Z | 3683 | hCoV-19/Luxembourg/LNS4184137/2020\|EPI_ISL_740430\| | ···················· | ···················· | ···················· | ···················· | ···················· | ···················· | | |

```
Z    3684 hCoV-19/Luxembourg/LNS4210904/2020|EPI_ISL_740431|    ····················  ····················  ····················  ····················  ·····T··············  ····················
Z    3685 hCoV-19/Luxembourg/LNS4223389/2020|EPI_ISL_740432|    ····················  ····················  ····················  ····················  ····················  ····················
Z    3686 hCoV-19/Luxembourg/LNS4227937/2020|EPI_ISL_740433|    ····················  ····················  ····················  ····················  ····················  ····················
Z    3687 hCoV-19/Luxembourg/LNS4229406/2020|EPI_ISL_740434|    ····················  ····················  ····················  ····················  ····················  ····················
Z    3688 hCoV-19/Luxembourg/LNS4235848/2020|EPI_ISL_740436|    ····················  ····················  ····················  ····················  ····················  ····················
Z    3689 hCoV-19/Luxembourg/LNS4237810/2020|EPI_ISL_740437|    ····················  ····················  ····················  ····················  ····················  ····················
Z    3690 hCoV-19/Luxembourg/LNS4239887/2020|EPI_ISL_740438|    ····················  ····················  ····················  ····················  ····················  ····················
Z    3691 hCoV-19/Luxembourg/LNS4269918/2020|EPI_ISL_740441|    ····················  ····················  ····················  ····················  ····················  ····················
Z    3692 hCoV-19/Luxembourg/LNS4280461/2020|EPI_ISL_740442|    ····················  ····················  ····················  ····················  ····················  ····················
Z    3693 hCoV-19/Luxembourg/LNS4281216/2020|EPI_ISL_740443|    ····················  ····················  ····················  ····················  ····················  ····················
Z    3694 hCoV-19/Luxembourg/LNS4300818/2020|EPI_ISL_740444|    ····················  ····················  ····················  ····················  ····················  ····················
Z    3695 hCoV-19/Luxembourg/LNS4302819/2020|EPI_ISL_740445|    ····················  ····················  ····················  ····················  ····················  ····················
Z    3696 hCoV-19/Luxembourg/LNS4305742/2020|EPI_ISL_740446|    ····················  ····················  ····················  ····················  ····················  ····················
Z    3697 hCoV-19/Luxembourg/LNS4322964/2020|EPI_ISL_740447|    ····················  ····················  ····················  ····················  ····················  ····················
Z    3698 hCoV-19/Luxembourg/LNS4323813/2020|EPI_ISL_740448|    ····················  ····················  ····················  ····················  ····················  ····················
Z    3699 hCoV-19/Luxembourg/LNS4334876/2020|EPI_ISL_740449|    ····················  ····················  ····················  ····················  ····················  ····················
Z    3700 hCoV-19/Luxembourg/LNS4337676/2020|EPI_ISL_740450|    ····················  ····················  ····················  ····················  ····················  ····················
Z    3701 hCoV-19/Luxembourg/LNS4349266/2020|EPI_ISL_740451|    ····················  ····················  ····················  ····················  ····················  ····················
Z    3702 hCoV-19/Luxembourg/LNS4352628/2020|EPI_ISL_740452|    ····················  ····················  ····················  ····················  ····················  ····················
Z    3703 hCoV-19/Luxembourg/LNS4354558/2020|EPI_ISL_740453|    ····················  ····················  ····················  ····················  ····················  ····················
Z    3704 hCoV-19/Luxembourg/LNS4356762/2020|EPI_ISL_740454|    ····················  ····················  ····················  ····················  ····················  ····················
Z    3705 hCoV-19/Luxembourg/LNS4360137/2020|EPI_ISL_740455|    ····················  ····················  ····················  ····················  ····················  ····················
Z    3706 hCoV-19/Luxembourg/LNS4369621/2020|EPI_ISL_740456|    ····················  ····················  ····················  ····················  ····················  ····················
Z    3707 hCoV-19/Luxembourg/LNS4374495/2020|EPI_ISL_740457|    ····················  ····················  ····················  ····················  ····················  ····················
Z    3708 hCoV-19/Luxembourg/LNS4375344/2020|EPI_ISL_740458|    ····················  ····················  ····················  ····················  ····················  ····················
Z    3709 hCoV-19/Luxembourg/LNS4376605/2020|EPI_ISL_740459|    ····················  ····················  ····················  ····················  ····················  ····················
Z    3710 hCoV-19/Luxembourg/LNS4377160/2020|EPI_ISL_740460|    ····················  ····················  ····················  ····················  ····················  ····················
Z    3711 hCoV-19/Luxembourg/LNS4381225/2020|EPI_ISL_740461|    ····················  ····················  ····················  ····················  ····················  ····················
Z    3712 hCoV-19/Luxembourg/LNS4386977/2020|EPI_ISL_740462|    ····················  ····················  ····················  ····················  ····················  ····················
Z    3713 hCoV-19/Luxembourg/LNS4391569/2020|EPI_ISL_740463|    ····················  ····················  ····················  ····················  ····················  ····················
Z    3714 hCoV-19/Luxembourg/LNS4394462/2020|EPI_ISL_740464|    ····················  ····················  ····················  ····················  ····················  ····················
Z    3715 hCoV-19/England/PORT-2E926F/2020|EPI_ISL_741707|20    ····················  ····················  ····················  ····················  ·····T··············  ····················
Z    3716 hCoV-19/England/PORT-2E92D8/2020|EPI_ISL_741711|20    ····················  ····················  ····················  ····················  ·····T··············  ····················
Z    3717 hCoV-19/England/PORT-2EAB72/2020|EPI_ISL_741712|20    ····················  ····················  ····················  ····················  ····················  ····················
Z    3718 hCoV-19/England/PORT-2EABBE/2020|EPI_ISL_741714|20    ····················  ····················  ····················  ····················  ····················  ····················
Z    3719 hCoV-19/England/PORT-2EE622/2020|EPI_ISL_741717|20    ····················  ····················  ····················  ····················  ····················  ····················
Z    3720 hCoV-19/England/PORT-2EE631/2020|EPI_ISL_741718|20    ····················  ····················  ····················  ····················  ····················  ····················
Z    3721 hCoV-19/England/PORT-2EE640/2020|EPI_ISL_741719|20    ····················  ·A··················  ····················  ····················  ····················  ··A·················
Z    3722 hCoV-19/England/PORT-2EE65F/2020|EPI_ISL_741720|20    ····················  ····················  ····················  ····················  ····················  ····················
Z    3723 hCoV-19/England/PORT-2EE67D/2020|EPI_ISL_741721|20    ····················  ····················  ····················  ····················  ····················  ····················
Z    3724 hCoV-19/England/PORT-2EE69B/2020|EPI_ISL_741723|20    ····················  ····················  ····················  ····················  ····················  ····················
Z    3725 hCoV-19/England/PORT-2EE6AA/2020|EPI_ISL_741724|20    ····················  ···N················  ····················  ····················  ····················  ····················
Z    3726 hCoV-19/England/PORT-2EE6C8/2020|EPI_ISL_741726|20    ····················  ····················  ····················  ····················  ····················  ····················
Z    3727 hCoV-19/England/PORT-2F6047/2020|EPI_ISL_741730|20    ····················  ····················  ····················  ····················  ····················  ····················
Z    3728 hCoV-19/England/PORT-2F6065/2020|EPI_ISL_741732|20    ····················  ····················  ····················  ····················  ····················  ····················
Z    3729 hCoV-19/England/PORT-2F6074/2020|EPI_ISL_741733|20    ····················  ····················  ····················  ····················  ····················  ····················
Z    3730 hCoV-19/England/PORT-2F6092/2020|EPI_ISL_741734|20    ····················  ····················  ····················  ····················  ····················  ····················
Z    3731 hCoV-19/England/PORT-2F60A1/2020|EPI_ISL_741735|20    ····················  ····················  ····················  ····················  ····················  ····················
Z    3732 hCoV-19/England/PORT-2F60B0/2020|EPI_ISL_741736|20    ····················  ····················  ····················  ····················  ····················  ····················
Z    3733 hCoV-19/England/PORT-2F60CF/2020|EPI_ISL_741737|20    ····················  ····················  ····················  ····················  ····················  ····················
Z    3734 hCoV-19/England/PORT-2F6135/2020|EPI_ISL_741742|20    ····················  ····················  ····················  ····················  ····················  ····················
Z    3735 hCoV-19/England/PORT-2F6144/2020|EPI_ISL_741743|20    ····················  ····················  ····················  ····················  ····················  ····················
Z    3736 hCoV-19/England/PORT-2F6153/2020|EPI_ISL_741744|20    ····················  ····················  ····················  ····················  ····················  ····················
Z    3737 hCoV-19/England/PORT-2F6171/2020|EPI_ISL_741746|20    ····················  ····················  ····················  ····················  ·····T··············  ····················
Z    3738 hCoV-19/England/PORT-2F61AE/2020|EPI_ISL_741747|20    ····················  ····················  ····················  ····················  ····················  ····················
Z    3739 hCoV-19/England/PORT-2F61BD/2020|EPI_ISL_741748|20    ····················  ····················  ····················  ····················  ····················  ····················
Z    3740 hCoV-19/England/PORT-2F61EA/2020|EPI_ISL_741749|20    ····················  ····················  ····················  ····················  ····················  ····················
Z    3741 hCoV-19/England/PORT-2F6214/2020|EPI_ISL_741751|20    ····················  ····················  ····················  ····················  ····················  ····················
Z    3742 hCoV-19/England/PORT-2F6232/2020|EPI_ISL_741753|20    ····················  ····················  ····················  ····················  ····················  ····················
Z    3743 hCoV-19/England/PORT-2F629C/2020|EPI_ISL_741755|20    ····················  ····················  ····················  ····················  ····················  ····················
Z    3745 hCoV-19/England/QEUH-A325FD/2020|EPI_ISL_741760|20    ····················  ····················  ····················  ····················  ····················  ····················
Z    3746 hCoV-19/England/QEUH-A4AFF4/2020|EPI_ISL_741761|20    ····················  ····················  ····················  ····················  ····················  ····················
Z    3747 hCoV-19/England/QEUH-A4B95F/2020|EPI_ISL_741762|20    ····················  ····················  ····················  ····················  ····················  ····················
Z    3748 hCoV-19/England/QEUH-A4BB86/2020|EPI_ISL_741763|20    ····················  ····················  ····················  ····················  ····················  ····················
Z    3749 hCoV-19/England/QEUH-A4BEAB/2020|EPI_ISL_741764|20    ····················  ····················  ····················  ····················  ····················  ····················
Z    3750 hCoV-19/England/QEUH-A4D42F/2020|EPI_ISL_741765|20    ····················  ····················  ····················  ····················  ····················  ····················
Z    3751 hCoV-19/England/QEUH-A622DF/2020|EPI_ISL_741766|20    ····················  ····················  ····················  ····················  ····················  ····················
Z    3752 hCoV-19/England/QEUH-A62521/2020|EPI_ISL_741767|20    ····················  ····················  ····················  ····················  ····················  ····················
Z    3753 hCoV-19/England/QEUH-A6261F/2020|EPI_ISL_741768|20    ····················  ····················  ····················  ····················  ····················  ····················
Z    3754 hCoV-19/England/QEUH-A63EDA/2020|EPI_ISL_741769|20    ····················  ····················  ····················  ····················  ····················  ····················
Z    3755 hCoV-19/England/QEUH-A78494/2020|EPI_ISL_741770|20    ····················  ····················  ····················  ····················  ····················  ····················
Z    3756 hCoV-19/England/QEUH-A78722/2020|EPI_ISL_741771|20    ····················  ····················  ····················  ····················  ····················  ····················
Z    3757 hCoV-19/England/QEUH-AAD01A/2020|EPI_ISL_741773|20    ····················  ····················  ····················  ····················  ····················  ····················
Z    3758 hCoV-19/England/QEUH-AAE3E3/2020|EPI_ISL_741774|20    ····················  ····················  ····················  ····················  ····················  ····················
Z    3759 hCoV-19/England/QEUH-AAE3F2/2020|EPI_ISL_741775|20    ····················  ····················  ····················  ····················  ····················  ····················
Z    3760 hCoV-19/England/QEUH-AAF2F4/2020|EPI_ISL_741776|20    ····················  ····················  ····················  ····················  ····················  ····················
Z    3761 hCoV-19/England/QEUH-B7C462/2020|EPI_ISL_741777|20    ····················  ····················  ····················  ····················  ····················  ····················
Z    3762 hCoV-19/England/QEUH-B7C471/2020|EPI_ISL_741778|20    ····················  ····················  ····················  ····················  ····················  ····················
Z    3763 hCoV-19/England/QEUH-B7C49F/2020|EPI_ISL_741779|20    ····················  ····················  ····················  ····················  ····················  ····················
Z    3764 hCoV-19/England/QEUH-B7C4AE/2020|EPI_ISL_741780|20    ····················  ·A··················  ····················  ····················  ····················  ··A·················
```

| | | | | | | | | |
|---|---|---|---|---|---|---|---|---|
| Z | 3765 | hCoV-19/England/QEUH-B7C4CC/2020\|EPI_ISL_741781\|20 | ···················· | ···················· | ····················· | ···················· | ···················· | ···················· |
| Z | 3766 | hCoV-19/England/QEUH-B7C4EA/2020\|EPI_ISL_741782\|20 | ···················· | ···················· | ····················· | ···················· | ···················· | ···················· |
| Z | 3767 | hCoV-19/England/QEUH-B7C4F9/2020\|EPI_ISL_741783\|20 | ···················· | ···················· | ····················· | ···················· | ···················· | ···················· |
| Z | 3768 | hCoV-19/England/QEUH-B7C514/2020\|EPI_ISL_741784\|20 | ···················· | ···················· | ····················· | ···················· | ···················· | ···················· |
| Z | 3769 | hCoV-19/England/QEUH-B7C56F/2020\|EPI_ISL_741786\|20 | ···················· | ···················· | ····················· | ···················· | ···················· | ···················· |
| Z | 3770 | hCoV-19/England/QEUH-B7C57E/2020\|EPI_ISL_741787\|20 | ···················· | ···················· | ····················· | ···················· | ···················· | ···················· |
| Z | 3771 | hCoV-19/England/QEUH-B7C58D/2020\|EPI_ISL_741788\|20 | ···················· | ···················· | ····················· | ···················· | ···················· | ···················· |
| Z | 3772 | hCoV-19/England/QEUH-B7C5C9/2020\|EPI_ISL_741789\|20 | ···················· | ···················· | ····················· | ···················· | ···················· | ···················· |
| Z | 3773 | hCoV-19/England/QEUH-B7C5D8/2020\|EPI_ISL_741790\|20 | ···················· | ···················· | ····················· | ···················· | ···················· | ···················· |
| Z | 3774 | hCoV-19/England/QEUH-B7C5F6/2020\|EPI_ISL_741791\|20 | ···················· | ···················· | ····················· | ···················· | ···················· | ···················· |
| Z | 3775 | hCoV-19/England/QEUH-B7C602/2020\|EPI_ISL_741792\|20 | ···················· | ···················· | ····················· | ···················· | ···················· | ···················· |
| Z | 3776 | hCoV-19/England/QEUH-B7C839/2020\|EPI_ISL_741793\|20 | ···················· | ···················· | ····················· | ···················· | ···················· | ···················· |
| Z | 3777 | hCoV-19/England/QEUH-B7C857/2020\|EPI_ISL_741794\|20 | ···················· | ···················· | ····················· | ···················· | ···················· | ···················· |
| Z | 3778 | hCoV-19/England/QEUH-B7C884/2020\|EPI_ISL_741795\|20 | ···················· | ···················· | ····················· | ···················· | ···················· | ···················· |
| Z | 3779 | hCoV-19/England/QEUH-B7C893/2020\|EPI_ISL_741796\|20 | ···················· | ···················· | ····················· | ···················· | ···················· | ···················· |
| Z | 3780 | hCoV-19/England/QEUH-B7C8A2/2020\|EPI_ISL_741797\|20 | ···················· | ···················· | ····················· | ···················· | ···················· | ···················· |
| Z | 3781 | hCoV-19/Russia/MOS-CRIE-13351661/2020\|EPI_ISL_7452 | ···················· | ···················· | ····················· | ···················· | ···················· | ···················· |
| Z | 3782 | hCoV-19/Russia/MOS-CRIE-13381296/2020\|EPI_ISL_7452 | ···················· | ···················· | ····················· | ···················· | ···················· | ···················· |
| Z | 3783 | hCoV-19/Russia/MOS-CRIE-13389205/2020\|EPI_ISL_7452 | ···················· | ···················· | ····················· | ···················· | ···················· | ···················· |
| Z | 3784 | hCoV-19/Russia/MOS-CRIE-13472261/2020\|EPI_ISL_7452 | ···················· | ···················· | ····················· | ···················· | ···················· | ···················· |
| Z | 3785 | hCoV-19/Russia/MOS-CRIE-13649167/2020\|EPI_ISL_7452 | ···················· | ···················· | ····················· | ···················· | ···················· | ···················· |
| Z | 3786 | hCoV-19/Russia/MOS-CRIE-13678204/2020\|EPI_ISL_7452 | ···················· | ···················· | ····················· | ···················· | ···················· | ···················· |
| Z | 3787 | hCoV-19/Russia/MOS-CRIE-13676494/2020\|EPI_ISL_7452 | ···················· | ···················· | ····················· | ···················· | ···················· | ···················· |
| Z | 3788 | hCoV-19/Russia/MOS-CRIE-13680794/2020\|EPI_ISL_7452 | ···················· | ···················· | ····················· | ···················· | ···················· | ···················· |
| Z | 3789 | hCoV-19/Russia/MOS-CRIE-13675465/2020\|EPI_ISL_7452 | ···················· | ···················· | ····················· | ···················· | ···················· | ···················· |
| Z | 3790 | hCoV-19/Russia/MOS-CRIE-13681164/2020\|EPI_ISL_7452 | ···················· | ···················· | ·········T··········· | ···················· | ···················· | ···················· |
| Z | 3791 | hCoV-19/Russia/MOS-CRIE-13668879/2020\|EPI_ISL_7452 | ···················· | ···················· | ····················· | ···················· | ···················· | ···················· |
| Z | 3792 | hCoV-19/Russia/MOS-CRIE-13679342/2020\|EPI_ISL_7452 | ···················· | ···················· | ····················· | ···················· | ···················· | ···················· |
| Z | 3793 | hCoV-19/Russia/MOS-CRIE-13678497/2020\|EPI_ISL_7452 | ···················· | ···················· | ····················· | ···················· | ···················· | ···················· |
| Z | 3794 | hCoV-19/Russia/MOS-CRIE-13672306/2020\|EPI_ISL_7452 | ···················· | ···················· | ····················· | ···················· | ···················· | ···················· |
| Z | 3795 | hCoV-19/Russia/MOS-CRIE-13642350/2020\|EPI_ISL_7452 | ···················· | ···················· | ····················· | ···················· | ···················· | ···················· |
| Z | 3796 | hCoV-19/Russia/MOS-CRIE-13670631/2020\|EPI_ISL_7452 | ···················· | ···················· | ····················· | ···················· | ···················· | ···················· |
| Z | 3797 | hCoV-19/Russia/MOS-CRIE-13678675/2020\|EPI_ISL_7452 | ···················· | ···················· | ····················· | ···················· | ···················· | ···················· |
| Z | 3798 | hCoV-19/Russia/MOS-CRIE-13644986/2020\|EPI_ISL_7452 | ···················· | ···················· | ····················· | ···················· | ···················· | ···················· |
| Z | 3799 | hCoV-19/Russia/MOS-CRIE-13678038/2020\|EPI_ISL_7452 | ···················· | ···················· | ····················· | ···················· | ···················· | ···················· |
| Z | 3800 | hCoV-19/Russia/MOS-CRIE-7172114/2020\|EPI_ISL_74525 | ···················· | ···················· | ····················· | ···················· | ···················· | ···················· |
| Z | 3801 | hCoV-19/USA/TX-DSHS-1834/2020\|EPI_ISL_745280\|2020- | ···················· | ···················· | ····················· | ···················· | ···················· | ···················· |
| Z | 3802 | hCoV-19/USA/TX-DSHS-1836/2020\|EPI_ISL_745281\|2020- | ···················· | ···················· | ····················· | ···················· | ···················· | ···················· |
| Z | 3803 | hCoV-19/USA/TX-DSHS-1837/2020\|EPI_ISL_745282\|2020- | ···················· | ···················· | ····················· | ···················· | ···················· | ···················· |
| Z | 3804 | hCoV-19/USA/TX-DSHS-1839/2020\|EPI_ISL_745283\|2020- | ···················· | ···················· | ····················· | ···················· | ···················· | ···················· |
| Z | 3805 | hCoV-19/USA/TX-DSHS-1842/2020\|EPI_ISL_745284\|2020- | ···················· | ···················· | ····················· | ···················· | ···················· | ···················· |
| Z | 3806 | hCoV-19/USA/TX-DSHS-1844/2020\|EPI_ISL_745285\|2020- | ···················· | ···················· | ····················· | ···················· | ···················· | ···················· |
| Z | 3807 | hCoV-19/USA/TX-DSHS-1845/2020\|EPI_ISL_745286\|2020- | ···················· | ···················· | ····················· | ···················· | ···················· | ···················· |
| Z | 3808 | hCoV-19/USA/TX-DSHS-1848/2020\|EPI_ISL_745287\|2020- | ···················· | ···················· | ····················· | ···················· | ···················· | ···················· |
| Z | 3809 | hCoV-19/USA/TX-DSHS-1850/2020\|EPI_ISL_745288\|2020- | ···················· | ···················· | ····················· | ···················· | ···················· | ···················· |
| Z | 3810 | hCoV-19/USA/TX-DSHS-1851/2020\|EPI_ISL_745289\|2020- | ···················· | ···················· | ····················· | ···················· | ···················· | ···················· |
| Z | 3811 | hCoV-19/USA/TX-DSHS-1852/2020\|EPI_ISL_745290\|2020- | ···················· | ···················· | ····················· | ···················· | ···················· | ···················· |
| Z | 3812 | hCoV-19/USA/TX-DSHS-1854/2020\|EPI_ISL_745291\|2020- | ···················· | ···················· | ····················· | ···················· | ···················· | ···················· |
| Z | 3813 | hCoV-19/USA/TX-DSHS-1856/2020\|EPI_ISL_745292\|2020- | ···················· | ···················· | ····················· | ···················· | ···················· | ···················· |
| Z | 3814 | hCoV-19/USA/TX-DSHS-1857/2020\|EPI_ISL_745293\|2020- | ···················· | ···················· | ····················· | ···················· | ···················· | ···················· |
| Z | 3815 | hCoV-19/USA/TX-DSHS-1858/2020\|EPI_ISL_745294\|2020- | ···················· | ···················· | ····················· | ···················· | ···················· | ···················· |
| Z | 3816 | hCoV-19/USA/TX-DSHS-1859/2020\|EPI_ISL_745295\|2020- | ···················· | ···················· | ····················· | ···················· | ···················· | ···················· |
| Z | 3817 | hCoV-19/USA/TX-DSHS-1861/2020\|EPI_ISL_745296\|2020- | ···················· | ···················· | ····················· | ···················· | ···················· | ···················· |
| Z | 3818 | hCoV-19/USA/TX-DSHS-1862/2020\|EPI_ISL_745297\|2020- | ···················· | ·A·················· | ····················· | ···················· | ···················· | ···················· |
| Z | 3819 | hCoV-19/USA/TX-DSHS-1863/2020\|EPI_ISL_745298\|2020- | ···················· | ···················· | ····················· | ···················· | ···················· | ···················· |
| Z | 3820 | hCoV-19/USA/TX-DSHS-1864/2020\|EPI_ISL_745299\|2020- | ···················· | ···················· | ····················· | ···················· | ···················· | ···················· |
| Z | 3821 | hCoV-19/Chile/RM-110881/2020\|EPI_ISL_746570\|2020-0 | ···················· | ···················· | ····················· | ···················· | ···················· | ···················· |
| Z | 3822 | hCoV-19/Chile/RM-110884/2020\|EPI_ISL_746571\|2020-0 | ···················· | ···················· | ····················· | ···················· | ···················· | ···················· |
| Z | 3823 | hCoV-19/Chile/RM-110889/2020\|EPI_ISL_746572\|2020-0 | ···················· | ···················· | ····················· | ···················· | ···················· | ···················· |
| Z | 3824 | hCoV-19/Chile/RM-110918/2020\|EPI_ISL_746573\|2020-0 | ···················· | ···················· | ····················· | ···················· | ···················· | ···················· |
| Z | 3825 | hCoV-19/Chile/RM-110919/2020\|EPI_ISL_746574\|2020-0 | ···················· | ···················· | ····················· | ···················· | ···················· | ···················· |
| Z | 3826 | hCoV-19/Chile/RM-110962/2020\|EPI_ISL_746575\|2020-0 | ···················· | ···················· | ····················· | ···················· | ···················· | ···················· |
| Z | 3827 | hCoV-19/Chile/RM-110966/2020\|EPI_ISL_746576\|2020-0 | ···················· | ···················· | ····················· | ···················· | ···················· | ···················· |
| Z | 3828 | hCoV-19/Chile/RM-110968/2020\|EPI_ISL_746577\|2020-0 | ···················· | ···················· | ····················· | ···················· | ···················· | ···················· |
| Z | 3829 | hCoV-19/Chile/RM-110981/2020\|EPI_ISL_746578\|2020-0 | ···················· | ···················· | ····················· | ···················· | ···················· | ···················· |
| Z | 3830 | hCoV-19/Chile/RM-110983/2020\|EPI_ISL_746579\|2020-0 | ···················· | ···················· | ····················· | ···················· | ···················· | ···················· |
| Z | 3831 | hCoV-19/Chile/RM-110987/2020\|EPI_ISL_746580\|2020-0 | ···················· | ···················· | ····················· | ···················· | ···················· | ···············A···· |
| Z | 3832 | hCoV-19/Chile/RM-111206/2020\|EPI_ISL_746581\|2020-0 | ···················· | ···················· | ····················· | ···················· | ···················· | ···················· |
| Z | 3833 | hCoV-19/Chile/RM-111211/2020\|EPI_ISL_746582\|2020-0 | ···················· | ···················· | ····················· | ···················· | ···················· | ···················· |
| Z | 3834 | hCoV-19/Chile/RM-111216/2020\|EPI_ISL_746583\|2020-0 | ···················· | ···················· | ····················· | ···················· | ···················· | ···················· |
| Z | 3835 | hCoV-19/Chile/RM-112466/2020\|EPI_ISL_746584\|2020-0 | ···················· | ···················· | ····················· | ···················· | ···················· | ···················· |
| Z | 3836 | hCoV-19/Chile/RM-112467/2020\|EPI_ISL_746585\|2020-0 | ···················· | ···················· | ····················· | ···················· | ···················· | ···················· |
| Z | 3837 | hCoV-19/Chile/RM-112634/2020\|EPI_ISL_746586\|2020-0 | ···················· | ···················· | ····················· | ···················· | ···················· | ···················· |
| Z | 3838 | hCoV-19/Chile/RM-112635/2020\|EPI_ISL_746587\|2020-0 | ···················· | ···················· | ····················· | ···················· | ···················· | ···················· |
| Z | 3839 | hCoV-19/Chile/RM-112637/2020\|EPI_ISL_746588\|2020-0 | ···················· | ···················· | ····················· | ···················· | ···················· | ···················· |
| Z | 3840 | hCoV-19/Chile/RM-112760/2020\|EPI_ISL_746589\|2020-0 | ···················· | ···················· | ····················· | ···················· | ···················· | ···················· |
| Z | 3842 | hCoV-19/USA/CA-CZB-14876/2020\|EPI_ISL_739441\|2020- | ···················· | ···················· | ····················· | ···················· | ···················· | ···················· |
| Z | 3843 | hCoV-19/USA/CA-CZB-14757/2020\|EPI_ISL_739442\|2020- | ···················· | ·A·················· | ····················· | ···················· | ···················· | ···················· |
| Z | 3844 | hCoV-19/USA/CA-CZB-15122/2020\|EPI_ISL_739443\|2020- | ···················· | ·A·················· | ····················· | ···················· | ···················· | ···················· |
| Z | 3846 | hCoV-19/USA/CA-CZB-13714/2020\|EPI_ISL_739480\|2020- | ···················· | ···················· | ····················· | ···················· | ···················· | ···················· |

| | | | | | | | | |
|---|---|---|---|---|---|---|---|---|
| Z | 3848 | hCoV-19/USA/CA-CZB-14685/2020\|EPI_ISL_739482\|2020- | ···················· | ···················· | ···················· | ···················· | ···················· | ···················· |
| Z | 3850 | hCoV-19/USA/CA-CZB-14087/2020\|EPI_ISL_739484\|2020- | ···················· | ···················· | ···················· | ···················· | ···················· | ···················· |
| Z | 3851 | hCoV-19/USA/CA-CZB-15339/2020\|EPI_ISL_739485\|2020- | ···················· | ···················· | ···················· | ···················· | ···················· | ···················· |
| Z | 3852 | hCoV-19/USA/CA-CZB-14234/2020\|EPI_ISL_739486\|2020- | ···················· | ···················· | ···················· | ···················· | ···················· | ···················· |
| Z | 3853 | hCoV-19/USA/CA-CZB-15971/2020\|EPI_ISL_739487\|2020- | ···················· | ·A·················· | ···················· | ···················· | ···················· | ···················· |
| Z | 3854 | hCoV-19/USA/CA-CZB-15441/2020\|EPI_ISL_739488\|2020- | ···················· | ···················· | ···················· | ···················· | ···················· | ···················· |
| Z | 3855 | hCoV-19/USA/CA-CZB-14721/2020\|EPI_ISL_739489\|2020- | ···················· | ·A·················· | ···················· | ···················· | ···················· | ···················· |
| Z | 3856 | hCoV-19/USA/CA-CZB-13715/2020\|EPI_ISL_739490\|2020- | ···················· | ···················· | ···················· | ···················· | ···················· | ···················· |
| Z | 3857 | hCoV-19/USA/CA-CZB-15309/2020\|EPI_ISL_739491\|2020- | ···················· | ·A·················· | ···················· | ···················· | ···················· | ···················· |
| Z | 3858 | hCoV-19/USA/CA-CZB-15490/2020\|EPI_ISL_739492\|2020- | ···················· | ·A·················· | ···················· | ···················· | ···················· | ···················· |
| Z | 3859 | hCoV-19/USA/CA-CZB-14695/2020\|EPI_ISL_739493\|2020- | ···················· | ···················· | ···················· | ···················· | ···················· | ···················· |
| Z | 3860 | hCoV-19/USA/CA-CZB-14204/2020\|EPI_ISL_739494\|2020- | ···················· | ···················· | ···················· | ···················· | ···················· | ···················· |
| Z | 3862 | hCoV-19/USA/CA-CZB-15405/2020\|EPI_ISL_739496\|2020- | ···················· | ···················· | ···················· | ···················· | ···················· | ···················· |
| Z | 3863 | hCoV-19/USA/CA-CZB-14902/2020\|EPI_ISL_739497\|2020- | ···················· | ···················· | ···················· | ···················· | ···················· | ···················· |
| Z | 3864 | hCoV-19/USA/CA-CZB-15091/2020\|EPI_ISL_739498\|2020- | ···················· | ···················· | ···················· | ···················· | ···················· | ···················· |
| Z | 3865 | hCoV-19/USA/CA-CZB-14856/2020\|EPI_ISL_739499\|2020- | ···················· | ···················· | ···················· | ···················· | ···················· | ···················· |
| Z | 3866 | hCoV-19/Luxembourg/LNS4139779/2020\|EPI_ISL_740426\| | ···················· | ···················· | ···················· | ···················· | ···················· | ···················· |
| Z | 3867 | hCoV-19/Luxembourg/LNS4149976/2020\|EPI_ISL_740427\| | ···················· | G··················· | ···················· | ···················· | ···················· | ···················· |
| Z | 3868 | hCoV-19/Luxembourg/LNS4169338/2020\|EPI_ISL_740428\| | ···················· | ····A··············· | ···················· | ···················· | ···················· | ···················· |
| Z | 3869 | hCoV-19/Luxembourg/LNS4410164/2020\|EPI_ISL_740467\| | ···················· | ····A··············· | ···················· | ···················· | ···················· | ···················· |
| Z | 3870 | hCoV-19/Luxembourg/LNS4432996/2020\|EPI_ISL_740468\| | ···················· | ···················· | ···················· | ···················· | ···················· | ···················· |
| Z | 3871 | hCoV-19/Luxembourg/LNS4448393/2020\|EPI_ISL_740469\| | ···················· | ···················· | ···················· | ···················· | ···················· | ···················· |
| Z | 3872 | hCoV-19/Luxembourg/LNS4453170/2020\|EPI_ISL_740470\| | ···················· | ···················· | ···················· | ···················· | ···················· | ···················· |
| Z | 3873 | hCoV-19/Luxembourg/LNS4455115/2020\|EPI_ISL_740471\| | ···················· | ···················· | ···················· | ···················· | ···················· | ···················· |
| Z | 3874 | hCoV-19/Luxembourg/LNS4456584/2020\|EPI_ISL_740472\| | ···················· | ···················· | ···················· | ···················· | ···················· | ···················· |
| Z | 3875 | hCoV-19/Luxembourg/LNS4463577/2020\|EPI_ISL_740474\| | ···················· | ···················· | ···················· | ···················· | ···················· | ···················· |
| Z | 3876 | hCoV-19/Luxembourg/LNS4464917/2020\|EPI_ISL_740475\| | ···················· | ···················· | ···················· | ···················· | ···················· | ···················· |
| Z | 3877 | hCoV-19/Luxembourg/LNS4470088/2020\|EPI_ISL_740476\| | ···················· | G··················· | ···················· | ···················· | ···················· | ···················· |
| Z | 3878 | hCoV-19/Luxembourg/LNS4470394/2020\|EPI_ISL_740477\| | ···················· | ···················· | ···················· | ···················· | ···················· | ···················· |
| Z | 3879 | hCoV-19/Luxembourg/LNS4478742/2020\|EPI_ISL_740478\| | ···················· | ···················· | ···················· | ···················· | ···················· | ···················· |
| Z | 3880 | hCoV-19/Luxembourg/LNS4482275/2020\|EPI_ISL_740479\| | ···················· | ···················· | ···················· | ···················· | ···················· | ···················· |
| Z | 3881 | hCoV-19/Luxembourg/LNS4484698/2020\|EPI_ISL_740480\| | ···················· | ···················· | ···················· | ···················· | ···················· | ···················· |
| Z | 3882 | hCoV-19/Luxembourg/LNS4497782/2020\|EPI_ISL_740481\| | ···················· | ···················· | ···················· | ···················· | ···················· | ···················· |
| Z | 3883 | hCoV-19/Luxembourg/LNS4499016/2020\|EPI_ISL_740482\| | ···················· | ···················· | ···················· | ···················· | ···················· | ···················· |
| Z | 3884 | hCoV-19/Luxembourg/LNS4499598/2020\|EPI_ISL_740483\| | ···················· | ···················· | ···················· | ···················· | ···················· | ···················· |
| Z | 3885 | hCoV-19/Luxembourg/LNS4515413/2020\|EPI_ISL_740484\| | ···················· | ···················· | ···················· | ···················· | ···················· | ···················· |
| Z | 3886 | hCoV-19/Luxembourg/LNS4516866/2020\|EPI_ISL_740485\| | ···················· | ···················· | ···················· | ···················· | ······T············· | ···················· |
| Z | 3887 | hCoV-19/Luxembourg/LNS4517528/2020\|EPI_ISL_740486\| | ···················· | ···················· | ···················· | ···················· | ···················· | ···················· |
| Z | 3888 | hCoV-19/Luxembourg/LNS4519371/2020\|EPI_ISL_740488\| | ···················· | ···················· | ···················· | ···················· | ···················· | ···················· |
| Z | 3889 | hCoV-19/Luxembourg/LNS4522010/2020\|EPI_ISL_740489\| | ···················· | ···················· | ···················· | ···················· | ···················· | ···················· |
| Z | 3890 | hCoV-19/Luxembourg/LNS4523860/2020\|EPI_ISL_740490\| | ···················· | ···················· | ···················· | ···················· | ···················· | ···················· |
| Z | 3891 | hCoV-19/Luxembourg/LNS4526018/2020\|EPI_ISL_740491\| | ···················· | G··················· | ···················· | ···················· | ···················· | ···················· |
| Z | 3892 | hCoV-19/Luxembourg/LNS4526500/2020\|EPI_ISL_740492\| | ···················· | ···················· | ···················· | ···················· | ···················· | ···················· |
| Z | 3893 | hCoV-19/Luxembourg/LNS4547737/2020\|EPI_ISL_740494\| | ···················· | ···················· | ···················· | ···················· | ···················· | ···················· |
| Z | 3894 | hCoV-19/Luxembourg/LNS4560213/2020\|EPI_ISL_740495\| | ···················· | ···················· | ···················· | ···················· | ···················· | ···················· |
| Z | 3895 | hCoV-19/Luxembourg/LNS4560700/2020\|EPI_ISL_740496\| | ···················· | ···················· | ···················· | ···················· | ···················· | ···················· |
| Z | 3896 | hCoV-19/Luxembourg/LNS4563609/2020\|EPI_ISL_740497\| | ···················· | ···················· | ···················· | ···················· | ···················· | ···················· |
| Z | 3897 | hCoV-19/Luxembourg/LNS4573955/2020\|EPI_ISL_740498\| | ···················· | ···················· | ···················· | ···················· | ···················· | ···················· |
| Z | 3898 | hCoV-19/Luxembourg/LNS4574827/2020\|EPI_ISL_740499\| | ···················· | ···················· | ···················· | ···················· | ···················· | ···················· |
| Z | 3899 | hCoV-19/England/PORT-2F62E7/2020\|EPI_ISL_741758\|20 | ···················· | ···················· | ···················· | ···················· | ···················· | ···················· |
| Z | 3900 | hCoV-19/England/PORT-2F6302/2020\|EPI_ISL_741759\|20 | ···················· | ···················· | ···················· | ···················· | ···················· | ···················· |
| Z | 3901 | hCoV-19/England/QEUH-B7C8C0/2020\|EPI_ISL_741798\|20 | ···················· | ···················· | ···················· | ···················· | ···················· | ···················· |
| Z | 3902 | hCoV-19/England/QEUH-B7C8DF/2020\|EPI_ISL_741799\|20 | ···················· | ···················· | ···················· | ···················· | ···················· | ···················· |
| Z | 3904 | hCoV-19/Wales/PHWC-4A2C9F/2020\|EPI_ISL_744001\|2020 | ···················· | ···················· | ···················· | ···················· | ···················· | ···················· |
| Z | 3905 | hCoV-19/Wales/PHWC-4A2CAE/2020\|EPI_ISL_744002\|2020 | ···················· | ···················· | ···················· | ···················· | ···················· | ···················· |
| Z | 3907 | hCoV-19/Wales/PHWC-4A2CCC/2020\|EPI_ISL_744004\|2020 | ···················· | ···················· | ···················· | ···················· | ···················· | ···················· |
| Z | 3908 | hCoV-19/Wales/PHWC-4A2CDB/2020\|EPI_ISL_744005\|2020 | ···················· | ···················· | ···················· | ···················· | ···················· | ···················· |
| Z | 3909 | hCoV-19/Wales/PHWC-4A2CEA/2020\|EPI_ISL_744006\|2020 | ···················· | ···················· | ···················· | ···················· | ···················· | ···················· |
| Z | 3910 | hCoV-19/Wales/PHWC-4A2CF9/2020\|EPI_ISL_744007\|2020 | ···················· | ···················· | ···················· | ···················· | ···················· | ···················· |
| Z | 3911 | hCoV-19/Wales/PHWC-4A2D23/2020\|EPI_ISL_744009\|2020 | ···················· | ···················· | ···················· | ···················· | ···················· | ···················· |
| Z | 3912 | hCoV-19/Wales/PHWC-4A2D41/2020\|EPI_ISL_744010\|2020 | ···················· | ···················· | ···················· | ···················· | ···················· | ···················· |
| Z | 3913 | hCoV-19/Wales/PHWC-4A2D50/2020\|EPI_ISL_744011\|2020 | ···················· | ···················· | ···················· | ···················· | ···················· | ···················· |
| Z | 3914 | hCoV-19/Wales/PHWC-4A2D6F/2020\|EPI_ISL_744012\|2020 | ···················· | ···················· | ···················· | ···················· | ···················· | ···················· |
| Z | 3915 | hCoV-19/Wales/PHWC-4A2D7E/2020\|EPI_ISL_744013\|2020 | ···················· | ···················· | ···················· | ···················· | ···················· | ···················· |
| Z | 3916 | hCoV-19/Wales/PHWC-4A2D8D/2020\|EPI_ISL_744014\|2020 | ···················· | ···················· | ···················· | ···················· | ···················· | ···················· |
| Z | 3917 | hCoV-19/Wales/PHWC-4A2D9C/2020\|EPI_ISL_744015\|2020 | ···················· | ···················· | ···················· | ···················· | ···················· | ···················· |
| Z | 3919 | hCoV-19/Wales/PHWC-4A2DC9/2020\|EPI_ISL_744017\|2020 | ···················· | ···················· | ···················· | ···················· | ···················· | ···················· |
| Z | 3921 | hCoV-19/Wales/PHWC-4A2DE7/2020\|EPI_ISL_744019\|2020 | ···················· | ···················· | ···················· | ···················· | ···················· | ···················· |
| Z | 3923 | hCoV-19/Wales/PHWC-4A2E02/2020\|EPI_ISL_744021\|2020 | ···················· | ···················· | ···················· | ···················· | ···················· | ···················· |
| Z | 3926 | hCoV-19/Wales/PHWC-4A2E4E/2020\|EPI_ISL_744024\|2020 | ···················· | ···················· | ···················· | ···················· | ···················· | ···················· |
| Z | 3927 | hCoV-19/Wales/PHWC-4A2E5D/2020\|EPI_ISL_744025\|2020 | ···················· | ···················· | ···················· | ···················· | ···················· | ···················· |
| Z | 3928 | hCoV-19/Wales/PHWC-4A2E6C/2020\|EPI_ISL_744026\|2020 | ···················· | ···················· | ···················· | ···················· | ···················· | ···················· |
| Z | 3929 | hCoV-19/Wales/PHWC-4A2E7B/2020\|EPI_ISL_744027\|2020 | ···················· | ···················· | ···················· | ···················· | ···················· | ···················· |
| Z | 3930 | hCoV-19/Wales/PHWC-4A2E99/2020\|EPI_ISL_744028\|2020 | ···················· | ···················· | ···················· | ···················· | ···················· | ···················· |
| Z | 3931 | hCoV-19/Wales/PHWC-4A30DE/2020\|EPI_ISL_744051\|2020 | ···················· | ···················· | ···················· | ···················· | ···················· | ···················· |
| Z | 3932 | hCoV-19/Wales/PHWC-4A3126/2020\|EPI_ISL_744056\|2020 | ···················· | ···················· | ···················· | ···················· | ···················· | ···················· |
| Z | 3933 | hCoV-19/Wales/PHWC-4A3135/2020\|EPI_ISL_744057\|2020 | ···················· | ···················· | ···················· | ···················· | ···················· | ···················· |
| Z | 3934 | hCoV-19/Wales/PHWC-4A3144/2020\|EPI_ISL_744058\|2020 | ···················· | ···················· | ···················· | ···················· | ···················· | ···················· |
| Z | 3935 | hCoV-19/Wales/PHWC-4A3162/2020\|EPI_ISL_744060\|2020 | ···················· | ···················· | ···················· | ···················· | ···················· | ···················· |
| Z | 3936 | hCoV-19/Wales/PHWC-4A3180/2020\|EPI_ISL_744062\|2020 | ···················· | ···················· | ···················· | ···················· | ···················· | ···················· |

| | | | | | | | | |
|---|---|---|---|---|---|---|---|---|
| Z | 3937 | hCoV-19/Wales/PHWC-4A319F/2020\|EPI_ISL_744063\|2020 | ···················· | ···················· | ····················· | ···················· | ···················· | ···················· |
| Z | 3938 | hCoV-19/Wales/PHWC-4A31AE/2020\|EPI_ISL_744064\|2020 | ···················· | ···················· | ····················· | ···················· | ···················· | ···················· |
| Z | 3939 | hCoV-19/Wales/PHWC-4A31BD/2020\|EPI_ISL_744065\|2020 | ···················· | ···················· | ····················· | ···················· | ···················· | ···················· |
| Z | 3940 | hCoV-19/Wales/PHWC-4A31EA/2020\|EPI_ISL_744068\|2020 | ···················· | ···················· | ····················· | ···················· | ···················· | ···················· |
| Z | 3942 | hCoV-19/USA/TX-DSHS-1865/2020\|EPI_ISL_745300\|2020- | ···················· | ····A··············· | ····················· | ···················· | ···················· | ···················· |
| Z | 3943 | hCoV-19/USA/TX-DSHS-1866/2020\|EPI_ISL_745301\|2020- | ···················· | ···················· | ····················· | ···················· | ···················· | ···················· |
| Z | 3944 | hCoV-19/USA/TX-DSHS-1869/2020\|EPI_ISL_745302\|2020- | ···················· | ···················· | ····················· | ···················· | ···················· | ···················· |
| Z | 3945 | hCoV-19/USA/TX-DSHS-1870/2020\|EPI_ISL_745303\|2020- | ···················· | ···················· | ····················· | ···················· | ···················· | ···················· |
| Z | 3946 | hCoV-19/USA/TX-DSHS-1871/2020\|EPI_ISL_745304\|2020- | ···················· | ·A·················· | ····················· | ···················· | ···················· | ···················· |
| Z | 3947 | hCoV-19/USA/TX-DSHS-1875/2020\|EPI_ISL_745305\|2020- | ···················· | ···················· | ····················· | ···················· | ···················· | ···················· |
| Z | 3948 | hCoV-19/USA/TX-DSHS-1829/2020\|EPI_ISL_745306\|2020- | ···················· | ···················· | ····················· | ···················· | ···················· | ···················· |
| Z | 3949 | hCoV-19/France/ARA-120965/2020\|EPI_ISL_745311\|2020 | ···················· | ···················· | ····················· | ···················· | ···················· | ···················· |
| Z | 3950 | hCoV-19/France/ARA-SC713/2020\|EPI_ISL_745312\|2020- | ···················· | ···················· | ····················· | ···················· | ···················· | ···················· |
| Z | 3951 | hCoV-19/France/ARA-SC714/2020\|EPI_ISL_745313\|2020- | ···················· | ···················· | ····················· | ···················· | ···················· | ···················· |
| Z | 3952 | hCoV-19/France/ARA-SC716/2020\|EPI_ISL_745314\|2020- | ···················· | ···················· | ····················· | ···················· | ···················· | ···················· |
| Z | 3953 | hCoV-19/France/ARA-SC717/2020\|EPI_ISL_745315\|2020- | ···················· | ···················· | ····················· | ···················· | ···················· | ···················· |
| Z | 3954 | hCoV-19/France/ARA-112956/2020\|EPI_ISL_745316\|2020 | ···················· | ···················· | ····················· | ···················· | ···················· | ···················· |
| Z | 3955 | hCoV-19/France/ARA-113925/2020\|EPI_ISL_745317\|2020 | ···················· | ···················· | ····················· | ···················· | ···················· | ···················· |
| Z | 3956 | hCoV-19/France/ARA-115781/2020\|EPI_ISL_745319\|2020 | ···················· | ···················· | ····················· | ···················· | ···················· | ···················· |
| Z | 3957 | hCoV-19/France/ARA-114092/2020\|EPI_ISL_745326\|2020 | ···················· | ···················· | ····················· | ···················· | ···················· | ···················· |
| Z | 3958 | hCoV-19/France/ARA-11455/2020\|EPI_ISL_745327\|2020- | ···················· | ···················· | ····················· | ···················· | ···················· | ···················· |
| Z | 3959 | hCoV-19/France/ARA-114632/2020\|EPI_ISL_745328\|2020 | ···················· | ···················· | ····················· | ···················· | ···················· | ···················· |
| Z | 3960 | hCoV-19/France/ARA-114791/2020\|EPI_ISL_745329\|2020 | ···················· | ···················· | ····················· | ···················· | ···················· | ···················· |
| Z | 3961 | hCoV-19/France/ARA-117723/2020\|EPI_ISL_745330\|2020 | ···················· | ···················· | ····················· | ···················· | ···················· | ···················· |
| Z | 3962 | hCoV-19/France/ARA-118288/2020\|EPI_ISL_745331\|2020 | ···················· | ···················· | ····················· | ···················· | ···················· | ···················· |
| Z | 3963 | hCoV-19/France/ARA-119478/2020\|EPI_ISL_745332\|2020 | ···················· | ···················· | ····················· | ···················· | ···················· | ···················· |
| Z | 3964 | hCoV-19/France/ARA-121156/2020\|EPI_ISL_745333\|2020 | ···················· | ···················· | ····················· | ···················· | ···················· | ···················· |
| Z | 3965 | hCoV-19/France/ARA-121597/2020\|EPI_ISL_745334\|2020 | ···················· | ···················· | ····················· | ···················· | ···················· | ···················· |
| Z | 3966 | hCoV-19/France/ARA-SC715/2020\|EPI_ISL_745335\|2020- | ···················· | ···················· | ····················· | ···················· | ···················· | ···················· |
| Z | 3967 | hCoV-19/France/ARA-SC718/2020\|EPI_ISL_745336\|2020- | ···················· | ···················· | ····················· | ···················· | ···················· | ···················· |
| Z | 3968 | hCoV-19/France/ARA-112613/2020\|EPI_ISL_745337\|2020 | ···················· | ···················· | ····················· | ···················· | ···················· | ···················· |
| Z | 3969 | hCoV-19/France/ARA-112755/2020\|EPI_ISL_745338\|2020 | ···················· | ···················· | ····················· | ···················· | ···················· | ···················· |
| Z | 3970 | hCoV-19/France/ARA-112775/2020\|EPI_ISL_745339\|2020 | ···················· | ···················· | ····················· | ···················· | ···················· | ···················· |
| Z | 3971 | hCoV-19/France/ARA-112789/2020\|EPI_ISL_745340\|2020 | ···················· | ···················· | ····················· | ···················· | ···················· | ···················· |
| Z | 3972 | hCoV-19/France/ARA-113019/2020\|EPI_ISL_745341\|2020 | ···················· | ···················· | ····················· | ···················· | ···················· | ···················· |
| Z | 3973 | hCoV-19/France/ARA-113354/2020\|EPI_ISL_745342\|2020 | ···················· | ···················· | ····················· | ···················· | ···················· | ···················· |
| Z | 3974 | hCoV-19/France/ARA-113383/2020\|EPI_ISL_745343\|2020 | ···················· | ···················· | ····················· | ···················· | ···················· | ···················· |
| Z | 3975 | hCoV-19/France/ARA-113416/2020\|EPI_ISL_745344\|2020 | ···················· | ···················· | ····················· | ···················· | ···················· | ···················· |
| Z | 3976 | hCoV-19/France/ARA-113487/2020\|EPI_ISL_745345\|2020 | ···················· | ···················· | ····················· | ···················· | ···················· | ···················· |
| Z | 3977 | hCoV-19/France/ARA-113814/2020\|EPI_ISL_745346\|2020 | ···················· | ···················· | ····················· | ···················· | ···················· | ···················· |
| Z | 3978 | hCoV-19/France/ARA-114153/2020\|EPI_ISL_745347\|2020 | ···················· | ···················· | ····················· | ···················· | ···················· | ···················· |
| Z | 3979 | hCoV-19/France/ARA-114176/2020\|EPI_ISL_745348\|2020 | ···················· | ···················· | ····················· | ···················· | ···················· | ···················· |
| Z | 3980 | hCoV-19/France/ARA-114292/2020\|EPI_ISL_745349\|2020 | ···················· | ···················· | ····················· | ···················· | ···················· | ···················· |
| Z | 3981 | hCoV-19/France/ARA-114435/2020\|EPI_ISL_745350\|2020 | ···················· | ···················· | ····················· | ···················· | ···················· | ···················· |
| Z | 3982 | hCoV-19/France/ARA-114442/2020\|EPI_ISL_745351\|2020 | ···················· | ···················· | ····················· | ···················· | ···················· | ···················· |
| Z | 3983 | hCoV-19/France/ARA-114488/2020\|EPI_ISL_745352\|2020 | ···················· | ···················· | ····················· | ···················· | ···················· | ···················· |
| Z | 3984 | hCoV-19/France/ARA-114784/2020\|EPI_ISL_745353\|2020 | ···················· | ···················· | ····················· | ···················· | ···················· | ···················· |
| Z | 3985 | hCoV-19/France/ARA-114869/2020\|EPI_ISL_745354\|2020 | ···················· | ······A············· | ····················· | ···················· | ···················· | ···················· |
| Z | 3986 | hCoV-19/France/ARA-114893/2020\|EPI_ISL_745355\|2020 | ···················· | ···················· | ····················· | ···················· | ···················· | ···················· |
| Z | 3987 | hCoV-19/France/ARA-114897/2020\|EPI_ISL_745356\|2020 | ···················· | ···················· | ····················· | ···················· | ···················· | ···················· |
| Z | 3988 | hCoV-19/France/ARA-115468/2020\|EPI_ISL_745357\|2020 | ···················· | ···················· | ····················· | ···················· | ···················· | ···················· |
| Z | 3989 | hCoV-19/France/ARA-115476/2020\|EPI_ISL_745358\|2020 | ···················· | ···················· | ····················· | ···················· | ···················· | ···················· |
| Z | 3990 | hCoV-19/France/ARA-115486/2020\|EPI_ISL_745359\|2020 | ···················· | ···················· | ····················· | ···················· | ···················· | ···················· |
| Z | 3991 | hCoV-19/France/ARA-1186-49/2020\|EPI_ISL_745380\|202 | ···················· | ···················· | ····················· | ···················· | ···················· | ···················· |
| Z | 3992 | hCoV-19/France/ARA-118746/2020\|EPI_ISL_745381\|2020 | ···················· | ···················· | ····················· | ···················· | ···················· | ···················· |
| Z | 3993 | hCoV-19/France/ARA-118811/2020\|EPI_ISL_745382\|2020 | ···················· | ···················· | ····················· | ···················· | ···················· | ···················· |
| Z | 3994 | hCoV-19/France/ARA-119183/2020\|EPI_ISL_745385\|2020 | ···················· | ···················· | ····················· | ···················· | ···················· | ···················· |
| Z | 3995 | hCoV-19/France/ARA-119333/2020\|EPI_ISL_745386\|2020 | ···················· | ···················· | ····················· | ···················· | ···················· | ···················· |
| Z | 3996 | hCoV-19/France/ARA-119458/2020\|EPI_ISL_745387\|2020 | ···················· | ···················· | ····················· | ···················· | ···················· | ···················· |
| Z | 3997 | hCoV-19/France/ARA-119460/2020\|EPI_ISL_745388\|2020 | ···················· | ···················· | ····················· | ···················· | ···················· | ···················· |
| Z | 3998 | hCoV-19/France/ARA-120170/2020\|EPI_ISL_745389\|2020 | ···················· | ·A·················· | ····················· | ···················· | ···················· | ···················· |
| Z | 3999 | hCoV-19/France/ARA-120894/2020\|EPI_ISL_745390\|2020 | ···················· | ···················· | ····················· | ···················· | ···················· | ···················· |
| Z | 4000 | hCoV-19/France/ARA-121169/2020\|EPI_ISL_745391\|2020 | ···················· | ···················· | ····················· | ···················· | ···················· | ···················· |
| Z | 4001 | hCoV-19/France/ARA-121875/2020\|EPI_ISL_745392\|2020 | ···················· | ···················· | ····················· | ···················· | ···················· | ···················· |
| Z | 4002 | hCoV-19/France/ARA-122291/2020\|EPI_ISL_745393\|2020 | ···················· | ···················· | ····················· | ···················· | ···················· | ···················· |
| Z | 4003 | hCoV-19/France/ARA-122351/2020\|EPI_ISL_745394\|2020 | ···················· | ···················· | ····················· | ···················· | ···················· | ···················· |
| Z | 4004 | hCoV-19/France/ARA-57377/2020\|EPI_ISL_745395\|2020- | ···················· | ···················· | ····················· | ···················· | ···················· | ···················· |
| Z | 4005 | hCoV-19/France/ARA-113522/2020\|EPI_ISL_745396\|2020 | ···················· | ···················· | ····················· | ···················· | ···················· | ···················· |
| Z | 4006 | hCoV-19/France/ARA-117754/2020\|EPI_ISL_745397\|2020 | ···················· | ···················· | ····················· | ···················· | ···················· | ···················· |
| Z | 4007 | hCoV-19/Chile/RM-127525/2020\|EPI_ISL_746600\|2020-0 | ···················· | ···················· | ····················· | ···················· | ···················· | ···················· |
| Z | 4008 | hCoV-19/Chile/RM-127562/2020\|EPI_ISL_746601\|2020-0 | ···················· | ···················· | ····················· | ···················· | ···················· | ···················· |
| Z | 4009 | hCoV-19/Chile/RM-127563/2020\|EPI_ISL_746602\|2020-0 | ···················· | ···················· | ····················· | ···················· | ···················· | ···················· |
| Z | 4010 | hCoV-19/Chile/RM-127568/2020\|EPI_ISL_746603\|2020-0 | ···················· | ···················· | ····················· | ···················· | ···················· | ···················· |
| Z | 4011 | hCoV-19/Chile/RM-127588/2020\|EPI_ISL_746604\|2020-0 | ···················· | ···················· | ····················· | ···················· | ···················· | ···················· |
| Z | 4012 | hCoV-19/Chile/RM-127602/2020\|EPI_ISL_746605\|2020-0 | ···················· | ···················· | ····················· | ···················· | ···················· | ···················· |
| Z | 4013 | hCoV-19/Chile/RM-127633/2020\|EPI_ISL_746606\|2020-0 | ···················· | ···················· | ····················· | ···················· | ···················· | ···················· |
| Z | 4014 | hCoV-19/Chile/RM-127636/2020\|EPI_ISL_746607\|2020-0 | ···················· | ···················· | ····················· | ···················· | ···················· | ···················· |
| Z | 4015 | hCoV-19/Chile/RM-127644/2020\|EPI_ISL_746608\|2020-0 | ···················· | ···················· | ····················· | ···················· | ···················· | ···················· |
| Z | 4016 | hCoV-19/Chile/RM-127666/2020\|EPI_ISL_746609\|2020-0 | ···················· | ···················· | ····················· | ···················· | ···················· | ···················· |
| Z | 4017 | hCoV-19/Chile/RM-127822/2020\|EPI_ISL_746610\|2020-0 | ···················· | ···················· | ····················· | ···················· | ···················· | ···················· |

| | | | | | | | | | |
|---|---|---|---|---|---|---|---|---|---|
| Z | 4018 | hCoV-19/Chile/RM-127908/2020\|EPI_ISL_746611\|2020-0 | ···················· | ···················· | ···················· | ···················· | ···················· | ···················· | |
| Z | 4019 | hCoV-19/Chile/RM-127976/2020\|EPI_ISL_746612\|2020-0 | ···················· | ···················· | ···················· | ···················· | ···················· | ············G······N | 1 |
| Z | 4020 | hCoV-19/Chile/RM-128039/2020\|EPI_ISL_746613\|2020-0 | ···················· | ···················· | ···················· | ···················· | ···················· | ···················· | |
| Z | 4021 | hCoV-19/Chile/RM-128080/2020\|EPI_ISL_746614\|2020-0 | ···················· | ···················· | ···················· | ···················· | ···················· | ···················· | |
| Z | 4022 | hCoV-19/Chile/RM-128084/2020\|EPI_ISL_746615\|2020-0 | ···················· | ···················· | ···················· | ···················· | ···················· | ···················· | |
| Z | 4023 | hCoV-19/Chile/RM-128176/2020\|EPI_ISL_746616\|2020-0 | ···················· | ···················· | ···················· | ···················· | ···················· | ···················· | |
| Z | 4024 | hCoV-19/Chile/RM-128225/2020\|EPI_ISL_746617\|2020-0 | ···················· | ···················· | ···················· | ···················· | ···················· | ···················· | |
| Z | 4025 | hCoV-19/Chile/RM-128269/2020\|EPI_ISL_746618\|2020-0 | ···················· | ···················· | ···················· | ···················· | ···················· | ···················· | |
| Z | 4026 | hCoV-19/Chile/RM-128531/2020\|EPI_ISL_746619\|2020-0 | ···················· | ···················· | ···················· | ···················· | ···················· | ···················· | |
| Z | 4027 | hCoV-19/Chile/RM-130072/2020\|EPI_ISL_746620\|2020-0 | ···················· | ···················· | ···················· | ···················· | ···················· | ···················· | |
| Z | 4028 | hCoV-19/Chile/RM-130073/2020\|EPI_ISL_746621\|2020-0 | ···················· | ···················· | ···················· | ···················· | ···················· | ···················· | |
| Z | 4029 | hCoV-19/Chile/RM-130081/2020\|EPI_ISL_746622\|2020-0 | ···················· | ···················· | ···················· | ···················· | ···················· | ···················· | |
| Z | 4030 | hCoV-19/Chile/RM-130924/2020\|EPI_ISL_746623\|2020-0 | ···················· | ···················· | ···················· | ···················· | ···················· | ···················· | |
| Z | 4031 | hCoV-19/Chile/RM-131047/2020\|EPI_ISL_746624\|2020-0 | ···················· | ···················· | ···················· | ···················· | ···················· | ···················· | |
| Z | 4032 | hCoV-19/Chile/RM-131381/2020\|EPI_ISL_746625\|2020-0 | ···················· | ···················· | ···················· | ···················· | ···················· | ···················· | |
| Z | 4033 | hCoV-19/Chile/RM-131430/2020\|EPI_ISL_746626\|2020-0 | ···················· | ···················· | ···················· | ···················· | ···················· | ···················· | |
| Z | 4034 | hCoV-19/Chile/RM-131476/2020\|EPI_ISL_746627\|2020-0 | ···················· | ···················· | ···················· | ···················· | ···················· | ···················· | |
| Z | 4035 | hCoV-19/Chile/RM-132398/2020\|EPI_ISL_746628\|2020-0 | ···················· | ···················· | ···················· | ···················· | ···················· | ···················· | |
| Z | 4036 | hCoV-19/Chile/RM-133091/2020\|EPI_ISL_746629\|2020-0 | ···················· | ···················· | ···················· | ···················· | ···················· | ···················· | |
| Z | 4037 | hCoV-19/Chile/RM-133092/2020\|EPI_ISL_746630\|2020-0 | ···················· | ···················· | ···················· | ···················· | ···················· | ···················· | |
| Z | 4038 | hCoV-19/Chile/RM-133147/2020\|EPI_ISL_746631\|2020-0 | ···················· | ···················· | ···················· | ···················· | ···················· | ···················· | |
| Z | 4039 | hCoV-19/Chile/RM-133191/2020\|EPI_ISL_746632\|2020-0 | ···················· | ···················· | ···················· | ···················· | ···················· | ···················· | |
| Z | 4040 | hCoV-19/Chile/RM-133240/2020\|EPI_ISL_746633\|2020-0 | ···················· | ···················· | ···················· | ···················· | ···················· | ···················· | |
| Z | 4041 | hCoV-19/Chile/RM-133318/2020\|EPI_ISL_746634\|2020-0 | ···················· | ···················· | ···················· | ···················· | ···················· | ···················· | |
| Z | 4042 | hCoV-19/Chile/RM-133547/2020\|EPI_ISL_746635\|2020-0 | ···················· | ···················· | ···················· | ···················· | ···················· | ···················· | |
| Z | 4043 | hCoV-19/Chile/RM-133558/2020\|EPI_ISL_746636\|2020-0 | ···················· | ···················· | ···················· | ···················· | ···················· | ···················· | |
| Z | 4044 | hCoV-19/Chile/RM-133562/2020\|EPI_ISL_746637\|2020-0 | ···················· | ···················· | ···················· | ···················· | ···················· | ···················· | |
| Z | 4045 | hCoV-19/Chile/RM-133625/2020\|EPI_ISL_746638\|2020-0 | ···················· | ···················· | ···················· | ···················· | ···················· | ···················· | |
| Z | 4046 | hCoV-19/Chile/RM-133627/2020\|EPI_ISL_746639\|2020-0 | ···················· | ···················· | ···················· | ···················· | ···················· | ···················· | |
| Z | 4047 | hCoV-19/Chile/RM-133746/2020\|EPI_ISL_746640\|2020-0 | ···················· | ···················· | ···················· | ···················· | ···················· | ···················· | |
| Z | 4048 | hCoV-19/Chile/RM-133831/2020\|EPI_ISL_746641\|2020-0 | ···················· | ···················· | ···················· | ···················· | ···················· | ···············A···· | |
| Z | 4049 | hCoV-19/Chile/RM-134034/2020\|EPI_ISL_746642\|2020-0 | ···················· | ···················· | ···················· | ···················· | ···················· | ···················· | |
| Z | 4050 | hCoV-19/Chile/RM-134067/2020\|EPI_ISL_746643\|2020-0 | ···················· | ···················· | ···················· | ···················· | ···················· | ···················· | |
| Z | 4051 | hCoV-19/Chile/RM-145979/2020\|EPI_ISL_746644\|2020-0 | ···················· | ···················· | ···················· | ···················· | ···················· | ···················· | |
| Z | 4052 | hCoV-19/Chile/RM-146431/2020\|EPI_ISL_746645\|2020-0 | ···················· | ···················· | ···················· | ···················· | ···················· | ···················· | |
| Z | 4053 | hCoV-19/Chile/RM-146538/2020\|EPI_ISL_746646\|2020-0 | ···················· | ···················· | ···················· | ···················· | ···················· | ···················· | |
| Z | 4054 | hCoV-19/Chile/RM-150435/2020\|EPI_ISL_746647\|2020-0 | ···················· | ···················· | ···················· | ···················· | ···················· | ···················· | |
| Z | 4055 | hCoV-19/Chile/RM-152036/2020\|EPI_ISL_746648\|2020-0 | ···················· | ···················· | ···················· | ···················· | ···················· | ···················· | |
| Z | 4056 | hCoV-19/Chile/RM-152037/2020\|EPI_ISL_746649\|2020-0 | ···················· | ···················· | ···················· | ···················· | ···················· | ···················· | |
| Z | 4057 | hCoV-19/Chile/RM-153333/2020\|EPI_ISL_746650\|2020-0 | ···················· | ···················· | ···················· | ···················· | ···················· | ···················· | |
| Z | 4058 | hCoV-19/Chile/RM-153360/2020\|EPI_ISL_746651\|2020-0 | ···················· | ·A·················· | ···················· | ···················· | ···················· | ···················· | |
| Z | 4059 | hCoV-19/Chile/RM-153390/2020\|EPI_ISL_746652\|2020-0 | ···················· | ···················· | ···················· | ···················· | ···················· | ···················· | |
| Z | 4060 | hCoV-19/Chile/RM-153914/2020\|EPI_ISL_746653\|2020-0 | ···················· | ···················· | ···················· | ···················· | ···················· | ···················· | |
| Z | 4061 | hCoV-19/Chile/RM-153915/2020\|EPI_ISL_746654\|2020-0 | ···················· | ·A·················· | ···················· | ···················· | ···················· | ···················· | |
| Z | 4062 | hCoV-19/Chile/RM-154196/2020\|EPI_ISL_746655\|2020-0 | ···················· | ···················· | ···················· | ···················· | ···················· | ···················· | |
| Z | 4063 | hCoV-19/Chile/RM-154362/2020\|EPI_ISL_746656\|2020-0 | ···················· | ···················· | ···················· | ···················· | ···················· | ···················· | |
| Z | 4064 | hCoV-19/Chile/RM-154655/2020\|EPI_ISL_746657\|2020-0 | ···················· | ···················· | ···················· | ···················· | ···················· | ···················· | |
| Z | 4065 | hCoV-19/Chile/RM-154667/2020\|EPI_ISL_746658\|2020-0 | ···················· | ···················· | ···················· | ···················· | ···················· | ···················· | |
| Z | 4066 | hCoV-19/Chile/RM-155380/2020\|EPI_ISL_746659\|2020-0 | ···················· | ···················· | ···················· | ···················· | ···················· | ···················· | |
| Z | 4067 | hCoV-19/Chile/RM-155487/2020\|EPI_ISL_746660\|2020-0 | ···················· | ···················· | ···················· | ···················· | ···················· | ···················· | |
| Z | 4068 | hCoV-19/Chile/RM-155633/2020\|EPI_ISL_746661\|2020-0 | ···················· | ···················· | ···················· | ···················· | ···················· | ···················· | |
| Z | 4069 | hCoV-19/Chile/RM-155699/2020\|EPI_ISL_746662\|2020-0 | ···················· | ···················· | ···················· | ···················· | ···················· | ···················· | |
| Z | 4070 | hCoV-19/Chile/RM-155723/2020\|EPI_ISL_746663\|2020-0 | ···················· | ···················· | ···················· | ···················· | ···················· | ···················· | |
| Z | 4071 | hCoV-19/Chile/RM-155909/2020\|EPI_ISL_746664\|2020-0 | ···················· | ···················· | ···················· | ···················· | ···················· | ···················· | |
| Z | 4072 | hCoV-19/Chile/RM-155990/2020\|EPI_ISL_746665\|2020-0 | ···················· | ···················· | ···················· | ···················· | ···················· | ···················· | |
| Z | 4073 | hCoV-19/Chile/RM-156030/2020\|EPI_ISL_746666\|2020-0 | ···················· | ···················· | ···················· | ···················· | ···················· | ···················· | |
| Z | 4074 | hCoV-19/Chile/RM-156453/2020\|EPI_ISL_746667\|2020-0 | ···················· | ···················· | ···················· | ···················· | ···················· | ···················· | |
| Z | 4075 | hCoV-19/Chile/RM-156567/2020\|EPI_ISL_746668\|2020-0 | ···················· | ···················· | ···················· | ···················· | ···················· | ···················· | |
| Z | 4076 | hCoV-19/Chile/RM-156709/2020\|EPI_ISL_746669\|2020-0 | ···················· | ···················· | ···················· | ···················· | ···················· | ···················· | |
| Z | 4077 | hCoV-19/Chile/RM-157016/2020\|EPI_ISL_746670\|2020-0 | ···················· | ···················· | ···················· | ···················· | ···················· | ···················· | |
| Z | 4078 | hCoV-19/Chile/RM-157134/2020\|EPI_ISL_746671\|2020-0 | ···················· | ···················· | ···················· | ···················· | ···················· | ···················· | |
| Z | 4079 | hCoV-19/Chile/RM-157305/2020\|EPI_ISL_746672\|2020-0 | ···················· | ···················· | ···················· | ···················· | ···················· | ···················· | |
| Z | 4080 | hCoV-19/Chile/RM-157307/2020\|EPI_ISL_746673\|2020-0 | ···················· | ···················· | ···················· | ···················· | ···················· | ···················· | |
| Z | 4081 | hCoV-19/Chile/RM-157308/2020\|EPI_ISL_746674\|2020-0 | ···················· | ···················· | ···················· | ···················· | ···················· | ···················· | |
| Z | 4082 | hCoV-19/Chile/RM-157480/2020\|EPI_ISL_746675\|2020-0 | ···················· | ···················· | ···················· | ···················· | ···················· | ···················· | |
| Z | 4083 | hCoV-19/Chile/RM-157618/2020\|EPI_ISL_746676\|2020-0 | ···················· | ···················· | ···················· | ···················· | ···················· | ···················· | |
| Z | 4084 | hCoV-19/Chile/RM-157632/2020\|EPI_ISL_746677\|2020-0 | ···················· | ···················· | ···················· | ···················· | ···················· | ···················· | |
| Z | 4085 | hCoV-19/Chile/RM-157643/2020\|EPI_ISL_746678\|2020-0 | ···················· | ···················· | ···················· | ···················· | ···················· | ···················· | |
| Z | 4086 | hCoV-19/Chile/RM-157658/2020\|EPI_ISL_746679\|2020-0 | ···················· | ···················· | ···················· | ···················· | ···················· | ···················· | |
| Z | 4087 | hCoV-19/Chile/RM-157770/2020\|EPI_ISL_746680\|2020-0 | ···················· | ···················· | ···················· | ···················· | ···················· | ···················· | |
| Z | 4088 | hCoV-19/Chile/RM-157771/2020\|EPI_ISL_746681\|2020-0 | ···················· | ···················· | ···················· | ···················· | ···················· | ···················· | |
| Z | 4089 | hCoV-19/Chile/RM-157801/2020\|EPI_ISL_746682\|2020-0 | ···················· | ···················· | ···················· | ···················· | ···················· | ···················· | |
| Z | 4090 | hCoV-19/Chile/RM-157806/2020\|EPI_ISL_746683\|2020-0 | ···················· | ···················· | ···················· | ···················· | ···················· | ···················· | |
| Z | 4091 | hCoV-19/Chile/RM-157813/2020\|EPI_ISL_746684\|2020-0 | ···················· | ···················· | ···················· | ···················· | ···················· | ···················· | |
| Z | 4092 | hCoV-19/Chile/RM-158079/2020\|EPI_ISL_746685\|2020-0 | ···················· | ···················· | ···················· | ···················· | ···················· | ···················· | |
| Z | 4093 | hCoV-19/Chile/RM-158209/2020\|EPI_ISL_746686\|2020-0 | ···················· | ···················· | ···················· | ···················· | ···················· | ···················· | |
| Z | 4094 | hCoV-19/Chile/RM-158239/2020\|EPI_ISL_746687\|2020-0 | ···················· | ···················· | ···················· | ···················· | ···················· | ···················· | |
| Z | 4095 | hCoV-19/Chile/RM-163679/2020\|EPI_ISL_746688\|2020-0 | ···················· | ···················· | ···················· | ···················· | ···················· | ···················· | |
| Z | 4096 | hCoV-19/Chile/RM-163995/2020\|EPI_ISL_746689\|2020-0 | ···················· | ···················· | ···················· | ···················· | ···················· | ···················· | |
| Z | 4097 | hCoV-19/USA/CA-CZB-15450/2020\|EPI_ISL_739500\|2020- | ···················· | ···················· | ···················· | ···················· | ···················· | ···················· | |

Z 4098 hCoV-19/USA/CA-CZB-15076/2020|EPI_ISL_739501|2020- ···················· ···················· ····················· ···················· ···················· ····················
Z 4100 hCoV-19/USA/CA-CZB-13968/2020|EPI_ISL_739503|2020- ···················· ···················· ····················· ···················· ···················· ····················
Z 4101 hCoV-19/USA/CA-CZB-13690/2020|EPI_ISL_739504|2020- ···················· ···················· ····················· ···················· ···················· ····················
Z 4102 hCoV-19/USA/CA-CZB-15933/2020|EPI_ISL_739505|2020- ···················· ···················· ····················· ···················· ···················· ····················
Z 4103 hCoV-19/USA/CA-CZB-13980/2020|EPI_ISL_739506|2020- ···················· ·A·················· ····················· ···················· ···················· ····················
Z 4104 hCoV-19/USA/CA-CZB-13748/2020|EPI_ISL_739507|2020- ···················· ···················· ····················· ···················· ···················· ····················
Z 4105 hCoV-19/USA/CA-CZB-15280/2020|EPI_ISL_739508|2020- ···················· ·A·················· ····················· ···················· ···················· ····················
Z 4106 hCoV-19/USA/CA-CZB-15172/2020|EPI_ISL_739509|2020- ···················· ·A·················· ····················· ···················· ···················· ····················
Z 4107 hCoV-19/USA/CA-CZB-15066/2020|EPI_ISL_739510|2020- ···················· ·A·················· ····················· ···················· ···················· ····················
Z 4108 hCoV-19/USA/CA-CZB-14098/2020|EPI_ISL_739511|2020- ···················· ···················· ····················· ···················· ···················· ····················
Z 4109 hCoV-19/USA/CA-CZB-15203/2020|EPI_ISL_739512|2020- ···················· ···················· ····················· ···················· ···················· ····················
Z 4110 hCoV-19/USA/CA-CZB-14896/2020|EPI_ISL_739513|2020- ···················· ···················· ····················· ···················· ···················· ····················
Z 4111 hCoV-19/USA/CA-CZB-11255/2020|EPI_ISL_739514|2020- ···················· ···················· ····················· ···················· ··········Y········· ····················
Z 4113 hCoV-19/USA/CA-CZB-14342/2020|EPI_ISL_739516|2020- ···················· ···················· ····················· ···················· ···················· ····················
Z 4114 hCoV-19/USA/CA-CZB-14884/2020|EPI_ISL_739517|2020- ···················· ···················· ····················· ···················· ···················· ····················
Z 4115 hCoV-19/USA/CA-CZB-13924/2020|EPI_ISL_739518|2020- ···················· ·A·················· ····················· ···················· ···················· ····················
Z 4116 hCoV-19/USA/CA-CZB-13920/2020|EPI_ISL_739519|2020- ···················· ·A·················· ····················· ···················· ···················· ····················
Z 4117 hCoV-19/USA/CA-CZB-15143/2020|EPI_ISL_739520|2020- ···················· ·A·················· ····················· ···················· ···················· ····················
Z 4121 hCoV-19/USA/CA-CZB-15400/2020|EPI_ISL_739524|2020- ···················· ···················· ····················· ···················· ···················· ····················
Z 4122 hCoV-19/USA/CA-CZB-14790/2020|EPI_ISL_739525|2020- ···················· ···················· ····················· ···················· ···················· ····················
Z 4123 hCoV-19/USA/CA-CZB-13854/2020|EPI_ISL_739526|2020- ···················· ·A·················· ····················· ···················· ···················· ····················
Z 4124 hCoV-19/USA/CA-CZB-14775/2020|EPI_ISL_739527|2020- ···················· ···················· ····················· ···················· ···················· ····················
Z 4127 hCoV-19/USA/CA-CZB-14756/2020|EPI_ISL_739530|2020- ···················· ···················· ····················· ···················· ···················· ····················
Z 4128 hCoV-19/USA/CA-CZB-14814/2020|EPI_ISL_739531|2020- ···················· ···················· ····················· ···················· ···················· ····················
Z 4129 hCoV-19/USA/CA-CZB-14649/2020|EPI_ISL_739532|2020- ···················· ···················· ····················· ···················· ···················· ····················
Z 4130 hCoV-19/USA/CA-CZB-14737/2020|EPI_ISL_739533|2020- ···················· ·A·················· ···········T········· ···················· ···················· ····················
Z 4131 hCoV-19/USA/CA-CZB-14696/2020|EPI_ISL_739534|2020- ···················· ···················· ····················· ···················· ···················· ····················
Z 4132 hCoV-19/USA/CA-CZB-13757/2020|EPI_ISL_739535|2020- ···················· ·A·················· ····················· ···················· ···················· ····················
Z 4133 hCoV-19/USA/CA-CZB-14511/2020|EPI_ISL_739536|2020- ···················· ·A·················· ····················· ···················· ···················· ····················
Z 4134 hCoV-19/USA/CA-CZB-15165/2020|EPI_ISL_739537|2020- ···················· ·A·················· ····················· ···················· ···················· ····················
Z 4135 hCoV-19/USA/CA-CZB-14099/2020|EPI_ISL_739538|2020- ···················· ···················· ····················· ···················· ···················· ····················
Z 4136 hCoV-19/USA/CA-CZB-14157/2020|EPI_ISL_739539|2020- ···················· ·A·················· ····················· ···················· ···················· ····················
Z 4137 hCoV-19/USA/CA-CZB-13718/2020|EPI_ISL_739540|2020- ···················· ···················· ····················· ···················· ···················· ····················
Z 4138 hCoV-19/USA/CA-CZB-14313/2020|EPI_ISL_739541|2020- ···················· ···················· ····················· ···················· ···················· ····················
Z 4139 hCoV-19/USA/CA-CZB-14796/2020|EPI_ISL_739542|2020- ···················· ···················· ····················· ···················· ···················· ····················
Z 4140 hCoV-19/USA/CA-CZB-15028/2020|EPI_ISL_739543|2020- ···················· ···················· ····················· ···················· ···················· ····················
Z 4141 hCoV-19/USA/CA-CZB-13945/2020|EPI_ISL_739544|2020- ···················· ···················· ····················· ···················· ···················· ····················
Z 4143 hCoV-19/USA/CA-CZB-14949/2020|EPI_ISL_739546|2020- ···················· ·A·················· ····················· ···················· ···················· ····················
Z 4144 hCoV-19/USA/CA-CZB-14668/2020|EPI_ISL_739547|2020- ···················· ···················· ····················· ···················· ···················· ····················
Z 4145 hCoV-19/USA/CA-CZB-15936/2020|EPI_ISL_739548|2020- ···················· ···················· ····················· ···················· ···················· ····················
Z 4146 hCoV-19/USA/CA-CZB-15084/2020|EPI_ISL_739549|2020- ···················· ···················· ····················· ···················· ···················· ····················
Z 4147 hCoV-19/USA/CA-CZB-15230/2020|EPI_ISL_739550|2020- ···················· ···················· ····················· ···················· ···················· ····················
Z 4148 hCoV-19/USA/CA-CZB-14052/2020|EPI_ISL_739551|2020- ···················· ···················· ····················· ···················· ···················· ····················
Z 4149 hCoV-19/USA/CA-CZB-14062/2020|EPI_ISL_739552|2020- ···················· ···················· ····················· ···················· ···················· ····················
Z 4150 hCoV-19/USA/CA-CZB-14786/2020|EPI_ISL_739553|2020- ···················· ·A·················· ····················· ···················· ···················· ····················
Z 4151 hCoV-19/USA/CA-CZB-15361/2020|EPI_ISL_739554|2020- ···················· ···················· ····················· ···················· ···················· ····················
Z 4153 hCoV-19/USA/CA-CZB-14885/2020|EPI_ISL_739556|2020- ···················· ···················· ····················· ···················· ···················· ····················
Z 4154 hCoV-19/USA/CA-CZB-11273/2020|EPI_ISL_739557|2020- ···················· ···················· ····················· ···················· ···················· ····················
Z 4155 hCoV-19/USA/CA-CZB-14743/2020|EPI_ISL_739558|2020- ···················· ·A·················· ···········T········· ···················· ···················· ····················
Z 4156 hCoV-19/USA/CA-CZB-14870/2020|EPI_ISL_739559|2020- ···················· ···················· ····················· ···················· ···················· ····················
Z 4158 hCoV-19/USA/CA-CZB-14637/2020|EPI_ISL_739591|2020- ···················· ···················· ····················· ···················· ···················· ····················
Z 4159 hCoV-19/USA/CA-CZB-15467/2020|EPI_ISL_739592|2020- ···················· ···················· ···T················· ···················· ···················· ····················
Z 4160 hCoV-19/USA/CA-CZB-15453/2020|EPI_ISL_739599|2020- ···················· ···················· ····················· ···················· ···················· ····················
Z 4161 hCoV-19/Luxembourg/LNS4577509/2020|EPI_ISL_740500| ···················· ···················· ····················· ···················· ···················· ····················
Z 4162 hCoV-19/Luxembourg/LNS4577991/2020|EPI_ISL_740501| ···················· ···················· ····················· ···················· ······T············· ····················
Z 4163 hCoV-19/Luxembourg/LNS4578556/2020|EPI_ISL_740502| ···················· ···················· ····················· ···················· ···················· ····················
Z 4164 hCoV-19/Luxembourg/LNS4595968/2020|EPI_ISL_740504| ···················· ···················· ····················· ···················· ···················· ····················
Z 4165 hCoV-19/Luxembourg/LNS4597411/2020|EPI_ISL_740505| ···················· ···················· ····················· ···················· ···················· ····················
Z 4166 hCoV-19/Luxembourg/LNS4605001/2020|EPI_ISL_740506| ···················· ···················· ····················· ···················· ···················· ····················
Z 4167 hCoV-19/Luxembourg/LNS4612109/2020|EPI_ISL_740507| ···················· ···················· ····················· ···················· ···················· ····················
Z 4168 hCoV-19/Luxembourg/LNS4612590/2020|EPI_ISL_740508| ···················· ···················· ····················· ···················· ···················· ····················
Z 4169 hCoV-19/Luxembourg/LNS4613975/2020|EPI_ISL_740509| ···················· ···················· ····················· ···················· ···················· ····················
Z 4170 hCoV-19/Luxembourg/LNS4614580/2020|EPI_ISL_740510| ···················· ···················· ····················· ···················· ···················· ····················
Z 4171 hCoV-19/Luxembourg/LNS4616804/2020|EPI_ISL_740511| ···················· ···················· ····················· ···················· ···················· ····················
Z 4172 hCoV-19/Luxembourg/LNS4619481/2020|EPI_ISL_740512| ···················· ···················· ····················· ···················· ···················· ····················
Z 4173 hCoV-19/Luxembourg/LNS4646138/2020|EPI_ISL_740513| ···················· ···················· ····················· ···················· ···················· ····················
Z 4174 hCoV-19/Luxembourg/LNS4647966/2020|EPI_ISL_740514| ···················· ···················· ····················· ···················· ···················· ····················
Z 4175 hCoV-19/Luxembourg/LNS4648424/2020|EPI_ISL_740515| ···················· ···················· ····················· ···················· ···················· ····················
Z 4176 hCoV-19/Luxembourg/LNS4650246/2020|EPI_ISL_740516| ···················· ···················· ····················· ···················· ······T············· ····················
Z 4177 hCoV-19/Luxembourg/LNS4658560/2020|EPI_ISL_740517| ···T················ ···················· ····················· ···················· ···················· ····················
Z 4178 hCoV-19/Luxembourg/LNS4661846/2020|EPI_ISL_740518| ···················· ···················· ····················· ···················· ···················· ····················
Z 4179 hCoV-19/Luxembourg/LNS4668283/2020|EPI_ISL_740519| ···················· ···················· ····················· ···················· ···················· ····················
Z 4180 hCoV-19/Luxembourg/LNS4668346/2020|EPI_ISL_740520| ···················· G··················· ····················· ···················· ···················· ····················
Z 4181 hCoV-19/Luxembourg/LNS4673059/2020|EPI_ISL_740521| ···················· ···················· ····················· ···················· ···················· ····················
Z 4182 hCoV-19/Luxembourg/LNS4676787/2020|EPI_ISL_740522| ···················· ···················· ····················· ···················· ···················· ····················
Z 4183 hCoV-19/Luxembourg/LNS4683076/2020|EPI_ISL_740523| ···················· ···················· ····················· ···················· ···················· ····················
Z 4184 hCoV-19/Luxembourg/LNS4684680/2020|EPI_ISL_740524| ···················· ···················· ····················· ···················· ···················· ····················
Z 4185 hCoV-19/Luxembourg/LNS4693548/2020|EPI_ISL_740525| ···················· ···················· ····················· ···················· ···················· ····················
Z 4186 hCoV-19/Luxembourg/LNS4695210/2020|EPI_ISL_740526| ···················· ···················· ····················· ···················· ···················· ····················
Z 4187 hCoV-19/Luxembourg/LNS4701310/2020|EPI_ISL_740527| ···················· ···················· ····················· ···················· ···················· ····················

| | | | | | | | | |
|---|---|---|---|---|---|---|---|---|
| Z | 4188 | hCoV-19/Luxembourg/LNS4701723/2020\|EPI_ISL_740528\| | ···················· | ···················· | ···················· | ···················· | ···················· | ···················· |
| Z | 4189 | hCoV-19/Luxembourg/LNS4715510/2020\|EPI_ISL_740529\| | ···················· | G··················· | ···················· | ···················· | ···················· | ···················· |
| Z | 4190 | hCoV-19/Luxembourg/LNS4718766/2020\|EPI_ISL_740530\| | ···················· | ···················· | ···················· | ···················· | ···················· | ···················· |
| Z | 4191 | hCoV-19/Luxembourg/LNS4723540/2020\|EPI_ISL_740531\| | ···················· | ···················· | ···················· | ···················· | ···················· | ···················· |
| Z | 4192 | hCoV-19/Luxembourg/LNS4727554/2020\|EPI_ISL_740532\| | ···················· | ···················· | ···················· | ···················· | ···················· | ···················· |
| Z | 4193 | hCoV-19/Luxembourg/LNS4729252/2020\|EPI_ISL_740533\| | ···················· | G··················· | ···················· | ···················· | ···················· | ···················· |
| Z | 4194 | hCoV-19/Luxembourg/LNS4733684/2020\|EPI_ISL_740534\| | ···················· | ···················· | ···················· | ···················· | ···················· | ···················· |
| Z | 4195 | hCoV-19/Luxembourg/LNS4736675/2020\|EPI_ISL_740535\| | ···················· | ···················· | ···················· | ···················· | ···················· | ···················· |
| Z | 4196 | hCoV-19/Luxembourg/LNS4737895/2020\|EPI_ISL_740536\| | ···················· | ···················· | ···················· | ···················· | ···················· | ···················· |
| Z | 4197 | hCoV-19/Luxembourg/LNS4739096/2020\|EPI_ISL_740537\| | ···················· | ···················· | ···················· | ···················· | ···················· | ···················· |
| Z | 4198 | hCoV-19/Luxembourg/LNS4751928/2020\|EPI_ISL_740538\| | ···················· | ···················· | ···················· | ···················· | ···················· | ···················· |
| Z | 4199 | hCoV-19/Luxembourg/LNS4755447/2020\|EPI_ISL_740539\| | ···················· | ···················· | ···················· | ···················· | ···················· | ···················· |
| Z | 4200 | hCoV-19/Luxembourg/LNS4766232/2020\|EPI_ISL_740540\| | ···················· | ···················· | ···················· | ···················· | ···················· | ···················· |
| Z | 4201 | hCoV-19/Luxembourg/LNS4770640/2020\|EPI_ISL_740541\| | ···················· | ···················· | ···················· | ···················· | ···················· | ···················· |
| Z | 4202 | hCoV-19/Luxembourg/LNS4778691/2020\|EPI_ISL_740542\| | ···················· | ···················· | ···················· | ···················· | ···················· | ···················· |
| Z | 4203 | hCoV-19/Luxembourg/LNS4778833/2020\|EPI_ISL_740543\| | ···················· | ···················· | ···················· | ···················· | ···················· | ···················· |
| Z | 4204 | hCoV-19/Canada/BC-BCCDC-6423/2020\|EPI_ISL_740577\|2 | ···················· | ·A·················· | ···················· | ···················· | ···················· | ···A················ |
| Z | 4205 | hCoV-19/Canada/BC-BCCDC-6425/2020\|EPI_ISL_740578\|2 | ···················· | ·A·················· | ···················· | ···················· | ···················· | ···A················ |
| Z | 4206 | hCoV-19/Canada/BC-BCCDC-6427/2020\|EPI_ISL_740580\|2 | ···················· | ·A·················· | ···················· | ···················· | ···················· | ···A················ |
| Z | 4207 | hCoV-19/Canada/BC-BCCDC-6428/2020\|EPI_ISL_740581\|2 | ···················· | ·A·················· | ···················· | ···················· | ···················· | ···A················ |
| Z | 4208 | hCoV-19/Canada/BC-BCCDC-6429/2020\|EPI_ISL_740582\|2 | ···················· | ·A·················· | ···················· | ···················· | ···················· | ···A················ |
| Z | 4209 | hCoV-19/Canada/BC-BCCDC-6430/2020\|EPI_ISL_740583\|2 | ···················· | ·A·················· | ···················· | ···················· | ···················· | ···A················ |
| Z | 4210 | hCoV-19/Canada/BC-BCCDC-6431/2020\|EPI_ISL_740584\|2 | ···················· | ·A·················· | ···················· | ···················· | ···················· | ···A················ |
| Z | 4211 | hCoV-19/Canada/BC-BCCDC-6432/2020\|EPI_ISL_740585\|2 | ···················· | ···················· | ···················· | ···················· | ···················· | ···················· |
| Z | 4212 | hCoV-19/Canada/BC-BCCDC-6433/2020\|EPI_ISL_740586\|2 | ···················· | ···················· | ···················· | ···················· | ···················· | ···················· |
| Z | 4213 | hCoV-19/Canada/BC-BCCDC-6435/2020\|EPI_ISL_740587\|2 | ···················· | ·A·················· | ···················· | ···················· | ···················· | ···A················ |
| Z | 4214 | hCoV-19/England/QEUH-B7C909/2020\|EPI_ISL_741800\|20 | ···················· | ···················· | ···················· | ···················· | ···················· | ···················· |
| Z | 4215 | hCoV-19/England/QEUH-B7C936/2020\|EPI_ISL_741801\|20 | ···················· | ···················· | ···················· | ···················· | ···················· | ···················· |
| Z | 4216 | hCoV-19/England/QEUH-B7C945/2020\|EPI_ISL_741802\|20 | ···················· | ···················· | ···················· | ···················· | ···················· | ···················· |
| Z | 4217 | hCoV-19/England/QEUH-B7C954/2020\|EPI_ISL_741803\|20 | ···················· | ···················· | ···················· | ···················· | ···················· | ···················· |
| Z | 4218 | hCoV-19/England/QEUH-B7C963/2020\|EPI_ISL_741804\|20 | ···················· | ···················· | ···················· | ···················· | ···················· | ···················· |
| Z | 4219 | hCoV-19/England/QEUH-B7C990/2020\|EPI_ISL_741806\|20 | ···················· | ···················· | ···················· | ···················· | ···················· | ···················· |
| Z | 4220 | hCoV-19/England/QEUH-B7CB8B/2020\|EPI_ISL_741807\|20 | ···················· | ·A·················· | ···················· | ···················· | ···················· | ···················· |
| Z | 4221 | hCoV-19/England/QEUH-B7CB9A/2020\|EPI_ISL_741808\|20 | ···················· | ·A·················· | ···················· | ···················· | ···················· | ···················· |
| Z | 4222 | hCoV-19/England/QEUH-B7CBB8/2020\|EPI_ISL_741809\|20 | ···················· | ···················· | ···················· | ···················· | ···················· | ···················· |
| Z | 4223 | hCoV-19/England/QEUH-B7CBC7/2020\|EPI_ISL_741810\|20 | ···················· | ···················· | ···················· | ···················· | ···················· | ···················· |
| Z | 4224 | hCoV-19/England/QEUH-B7CBE5/2020\|EPI_ISL_741811\|20 | ···················· | ···················· | ···················· | ···················· | ···················· | ···················· |
| Z | 4225 | hCoV-19/England/QEUH-B7CC00/2020\|EPI_ISL_741813\|20 | ···················· | ···················· | ···················· | ···················· | ···················· | ···················· |
| Z | 4226 | hCoV-19/England/QEUH-B7CC1F/2020\|EPI_ISL_741814\|20 | ···················· | ···················· | ···················· | ···················· | ···················· | ···················· |
| Z | 4227 | hCoV-19/England/QEUH-B7CC2E/2020\|EPI_ISL_741815\|20 | ···················· | ·A·················· | ···················· | ···················· | ···················· | ···················· |
| Z | 4228 | hCoV-19/England/QEUH-B7CC3D/2020\|EPI_ISL_741816\|20 | ···················· | ···················· | ···················· | ···················· | ···················· | ···················· |
| Z | 4229 | hCoV-19/England/QEUH-B7CC4C/2020\|EPI_ISL_741817\|20 | ···················· | ···················· | ···················· | ···················· | ···················· | ···················· |
| Z | 4230 | hCoV-19/England/QEUH-B7CC79/2020\|EPI_ISL_741818\|20 | ···················· | ···················· | ···················· | ···················· | ···················· | ···················· |
| Z | 4231 | hCoV-19/England/QEUH-B7CC88/2020\|EPI_ISL_741819\|20 | ···················· | ···················· | ···················· | ···················· | ···················· | ···················· |
| Z | 4232 | hCoV-19/England/QEUH-B7CC97/2020\|EPI_ISL_741820\|20 | ···················· | ···················· | ···················· | ···················· | ···················· | ···················· |
| Z | 4233 | hCoV-19/England/QEUH-B7CCD3/2020\|EPI_ISL_741821\|20 | ···················· | ···················· | ···················· | ···················· | ···················· | ···················· |
| Z | 4234 | hCoV-19/England/QEUH-B7CCE2/2020\|EPI_ISL_741822\|20 | ···················· | ···················· | ···················· | ···················· | ···················· | ···················· |
| Z | 4235 | hCoV-19/England/QEUH-B7CD2B/2020\|EPI_ISL_741823\|20 | ···················· | ···················· | ···················· | ···················· | ···················· | ···················· |
| Z | 4236 | hCoV-19/England/QEUH-B7CD49/2020\|EPI_ISL_741824\|20 | ···················· | ···················· | ···················· | ···················· | ···················· | ···················· |
| Z | 4237 | hCoV-19/England/QEUH-B7CD58/2020\|EPI_ISL_741825\|20 | ···················· | ···················· | ···················· | ···················· | ···················· | ···················· |
| Z | 4238 | hCoV-19/England/QEUH-B7CD67/2020\|EPI_ISL_741826\|20 | ···················· | ···················· | ···················· | ···················· | ······T············· | ···················· |
| Z | 4239 | hCoV-19/England/QEUH-B7CDC1/2020\|EPI_ISL_741827\|20 | ···················· | ···················· | ···················· | ···················· | ···················· | ···················· |
| Z | 4240 | hCoV-19/England/QEUH-B7CE91/2020\|EPI_ISL_741828\|20 | ···················· | ···················· | ···················· | ···················· | ···················· | ···················· |
| Z | 4241 | hCoV-19/England/QEUH-B7CEA0/2020\|EPI_ISL_741829\|20 | ···················· | ···················· | ···················· | ···················· | ···················· | ···················· |
| Z | 4242 | hCoV-19/England/QEUH-B7CEDD/2020\|EPI_ISL_741830\|20 | ···················· | ···················· | ···················· | ···················· | ···················· | ···················· |
| Z | 4243 | hCoV-19/England/QEUH-B7CF07/2020\|EPI_ISL_741831\|20 | ···················· | ···················· | ···················· | ···················· | ···················· | ···················· |
| Z | 4244 | hCoV-19/England/QEUH-B7CF16/2020\|EPI_ISL_741832\|20 | ···················· | ···················· | ···················· | ···················· | ···················· | ···················· |
| Z | 4245 | hCoV-19/England/QEUH-B7CF61/2020\|EPI_ISL_741833\|20 | ···················· | ···················· | ···················· | ···················· | ···················· | ···················· |
| Z | 4246 | hCoV-19/England/QEUH-B7CFCB/2020\|EPI_ISL_741835\|20 | ···················· | ···················· | ···················· | ···················· | ···················· | ···················· |
| Z | 4247 | hCoV-19/England/QEUH-B7CFDA/2020\|EPI_ISL_741836\|20 | ···················· | ···················· | ···················· | ···················· | ···················· | ···················· |
| Z | 4248 | hCoV-19/England/QEUH-B7CFF8/2020\|EPI_ISL_741838\|20 | ···················· | ···················· | ···················· | ···················· | ···················· | ···················· |
| Z | 4249 | hCoV-19/England/QEUH-B7D021/2020\|EPI_ISL_741840\|20 | ···················· | ···················· | ···················· | ···················· | ···················· | ···················· |
| Z | 4250 | hCoV-19/England/QEUH-B7D030/2020\|EPI_ISL_741841\|20 | ···················· | ···················· | ···················· | ···················· | ···················· | ···················· |
| Z | 4251 | hCoV-19/England/QEUH-B7D04F/2020\|EPI_ISL_741842\|20 | ···················· | ···················· | ···················· | ···················· | ···················· | ···················· |
| Z | 4252 | hCoV-19/England/QEUH-B7D07C/2020\|EPI_ISL_741843\|20 | ···················· | ···················· | ···················· | ···················· | ···················· | ···················· |
| Z | 4253 | hCoV-19/England/QEUH-B7D0C7/2020\|EPI_ISL_741844\|20 | ···················· | ···················· | ···················· | ···················· | ···················· | ···················· |
| Z | 4254 | hCoV-19/England/QEUH-B7D0D6/2020\|EPI_ISL_741845\|20 | ···················· | ···················· | ···················· | ···················· | ···················· | ···················· |
| Z | 4255 | hCoV-19/England/QEUH-B7D0E5/2020\|EPI_ISL_741846\|20 | ···················· | ···················· | ···················· | ···················· | ···················· | ···················· |
| Z | 4256 | hCoV-19/England/QEUH-B7D0F4/2020\|EPI_ISL_741847\|20 | ···················· | ···················· | ···················· | ···················· | ···················· | ···················· |
| Z | 4257 | hCoV-19/England/QEUH-B7D100/2020\|EPI_ISL_741848\|20 | ···················· | ···················· | ···················· | ···················· | ···················· | ···················· |
| Z | 4258 | hCoV-19/England/QEUH-B7D11F/2020\|EPI_ISL_741849\|20 | ···················· | ···················· | ···················· | ···················· | ···················· | ···················· |
| Z | 4259 | hCoV-19/England/QEUH-B7D2D0/2020\|EPI_ISL_741850\|20 | ···················· | ·A·················· | ···················· | ···················· | ···················· | ···A················ |
| Z | 4260 | hCoV-19/England/QEUH-B7D337/2020\|EPI_ISL_741851\|20 | ···················· | ···················· | ···················· | ···················· | ···················· | ···················· |
| Z | 4261 | hCoV-19/England/QEUH-B7D355/2020\|EPI_ISL_741852\|20 | ···················· | ···················· | ···················· | ···················· | ···················· | ···················· |
| Z | 4262 | hCoV-19/England/QEUH-B7D364/2020\|EPI_ISL_741853\|20 | ···················· | ···················· | ···················· | ···················· | ···················· | ···················· |
| Z | 4263 | hCoV-19/England/QEUH-B7D373/2020\|EPI_ISL_741854\|20 | ···················· | ·A·················· | ···················· | ···················· | ···················· | ···A················ |
| Z | 4264 | hCoV-19/England/QEUH-B7D391/2020\|EPI_ISL_741855\|20 | ···················· | ···················· | ···················· | ···················· | ···················· | ···················· |
| Z | 4265 | hCoV-19/England/QEUH-B7D3DD/2020\|EPI_ISL_741856\|20 | ···················· | ···················· | ···················· | ···················· | ···················· | ···················· |
| Z | 4266 | hCoV-19/England/QEUH-B7D3EC/2020\|EPI_ISL_741857\|20 | ···················· | ···················· | ···················· | ···················· | ···················· | ···················· |
| Z | 4267 | hCoV-19/England/QEUH-B7D3FB/2020\|EPI_ISL_741858\|20 | ···················· | ···················· | ···················· | ···················· | ···················· | ···················· |

| | | | | | | | | |
|---|---|---|---|---|---|---|---|---|
| Z | 4268 | hCoV-19/England/QEUH-B7D407/2020\|EPI_ISL_741859\|20 | .................... | .................... | .................... | .................... | .................... | .................... |
| Z | 4269 | hCoV-19/England/QEUH-B7D425/2020\|EPI_ISL_741860\|20 | .................... | .................... | .................... | .................... | .................... | .................... |
| Z | 4270 | hCoV-19/England/QEUH-B7D434/2020\|EPI_ISL_741861\|20 | .................... | .A.................. | .................... | .................... | .................... | ..A................. |
| Z | 4271 | hCoV-19/England/QEUH-B7D452/2020\|EPI_ISL_741862\|20 | .................... | .A.................. | .................... | .................... | .................... | ..A................. |
| Z | 4272 | hCoV-19/England/QEUH-B7D461/2020\|EPI_ISL_741863\|20 | .................... | .................... | .................... | .................... | .................... | .................... |
| Z | 4273 | hCoV-19/England/QEUH-B7D470/2020\|EPI_ISL_741864\|20 | .................... | .................... | .................... | .................... | .................... | .................... |
| Z | 4274 | hCoV-19/England/QEUH-B7D48F/2020\|EPI_ISL_741865\|20 | .................... | .................... | .................... | .................... | .................... | .................... |
| Z | 4275 | hCoV-19/England/QEUH-B7D4CB/2020\|EPI_ISL_741867\|20 | .................... | .................... | .................... | .................... | .................... | .................... |
| Z | 4276 | hCoV-19/England/QEUH-B7D4F8/2020\|EPI_ISL_741868\|20 | .................... | .................... | .................... | .................... | .................... | .................... |
| Z | 4277 | hCoV-19/England/QEUH-B7D504/2020\|EPI_ISL_741869\|20 | .................... | .................... | .................... | .................... | .................... | .................... |
| Z | 4278 | hCoV-19/England/QEUH-B7D513/2020\|EPI_ISL_741870\|20 | .................... | .................... | .................... | .................... | .................... | .................... |
| Z | 4279 | hCoV-19/England/QEUH-B7D531/2020\|EPI_ISL_741871\|20 | .................... | .................... | .................... | .................... | .................... | .................... |
| Z | 4280 | hCoV-19/England/QEUH-B7D540/2020\|EPI_ISL_741872\|20 | .................... | .................... | .................... | .................... | .................... | .................... |
| Z | 4281 | hCoV-19/England/QEUH-B7D58C/2020\|EPI_ISL_741873\|20 | .................... | .................... | .................... | .................... | .................... | .................... |
| Z | 4282 | hCoV-19/England/QEUH-B7D5AA/2020\|EPI_ISL_741874\|20 | .................... | .................... | .................... | .................... | .................... | .................... |
| Z | 4283 | hCoV-19/Wales/PHWC-4A2EB7/2020\|EPI_ISL_744030\|2020 | .................... | .................... | .................... | .................... | .................... | .................... |
| Z | 4285 | hCoV-19/Wales/PHWC-4A2F0F/2020\|EPI_ISL_744032\|2020 | .................... | .................... | .................... | .................... | .................... | .................... |
| Z | 4286 | hCoV-19/Wales/PHWC-4A2F3C/2020\|EPI_ISL_744034\|2020 | .................... | .................... | .................... | .................... | .................... | .................... |
| Z | 4287 | hCoV-19/Wales/PHWC-4A2F78/2020\|EPI_ISL_744037\|2020 | .................... | .................... | .................... | .................... | .................... | .................... |
| Z | 4288 | hCoV-19/Wales/PHWC-4A2F87/2020\|EPI_ISL_744038\|2020 | .................... | .................... | .................... | .................... | .................... | .................... |
| Z | 4291 | hCoV-19/Wales/PHWC-4A3029/2020\|EPI_ISL_744042\|2020 | .................... | .................... | .................... | .................... | .................... | .................... |
| Z | 4292 | hCoV-19/Wales/PHWC-4A3038/2020\|EPI_ISL_744043\|2020 | .................... | .................... | .................... | .................... | .................... | .................... |
| Z | 4293 | hCoV-19/Wales/PHWC-4A3047/2020\|EPI_ISL_744044\|2020 | .................... | .................... | .................... | .................... | .................... | .................... |
| Z | 4294 | hCoV-19/Wales/PHWC-4A3065/2020\|EPI_ISL_744045\|2020 | .................... | .................... | .................... | .................... | .................... | .................... |
| Z | 4295 | hCoV-19/Wales/PHWC-4A3074/2020\|EPI_ISL_744046\|2020 | .................... | .................... | .................... | .................... | .................... | .................... |
| Z | 4296 | hCoV-19/Wales/PHWC-4A3092/2020\|EPI_ISL_744047\|2020 | .................... | .................... | .................... | .................... | .................... | .................... |
| Z | 4297 | hCoV-19/Wales/PHWC-4A30A1/2020\|EPI_ISL_744048\|2020 | .................... | .................... | .................... | .................... | .................... | .................... |
| Z | 4298 | hCoV-19/Wales/PHWC-4A30B0/2020\|EPI_ISL_744049\|2020 | .................... | .................... | .................... | .................... | .................... | .................... |
| Z | 4299 | hCoV-19/Wales/PHWC-4A30ED/2020\|EPI_ISL_744052\|2020 | .................... | .................... | .................... | .................... | .................... | .................... |
| Z | 4300 | hCoV-19/Wales/PHWC-4A30FC/2020\|EPI_ISL_744053\|2020 | .................... | .................... | .................... | .................... | .................... | .................... |
| Z | 4301 | hCoV-19/Wales/PHWC-4A3117/2020\|EPI_ISL_744055\|2020 | .................... | .................... | .................... | .................... | .................... | .................... |
| Z | 4302 | hCoV-19/Wales/PHWC-4A3205/2020\|EPI_ISL_744070\|2020 | .................... | .................... | .................... | .................... | .................... | .................... |
| Z | 4303 | hCoV-19/Wales/PHWC-4A3232/2020\|EPI_ISL_744073\|2020 | .................... | .................... | .................... | .................... | .................... | .................... |
| Z | 4304 | hCoV-19/Wales/PHWC-4A3241/2020\|EPI_ISL_744074\|2020 | .................... | .................... | .................... | .................... | .................... | .................... |
| Z | 4305 | hCoV-19/Wales/PHWC-4A3250/2020\|EPI_ISL_744075\|2020 | .................... | .................... | .................... | .................... | .................... | .................... |
| Z | 4306 | hCoV-19/Wales/PHWC-4A326F/2020\|EPI_ISL_744076\|2020 | .................... | .................... | .................... | .................... | .................... | .................... |
| Z | 4307 | hCoV-19/Wales/PHWC-4A328D/2020\|EPI_ISL_744078\|2020 | .................... | .................... | .................... | .................... | .................... | .................... |
| Z | 4308 | hCoV-19/Wales/PHWC-4A329C/2020\|EPI_ISL_744079\|2020 | .................... | .................... | .................... | .................... | .................... | .................... |
| Z | 4309 | hCoV-19/Wales/PHWC-4A32AB/2020\|EPI_ISL_744080\|2020 | .................... | .................... | .................... | .................... | .................... | .................... |
| Z | 4311 | hCoV-19/Wales/PHWC-4A32D8/2020\|EPI_ISL_744083\|2020 | .................... | .................... | .................... | .................... | .................... | .................... |
| Z | 4312 | hCoV-19/Wales/PHWC-4A32F6/2020\|EPI_ISL_744085\|2020 | .................... | .................... | .................... | .................... | .................... | .................... |
| Z | 4314 | hCoV-19/Wales/PHWC-4A3311/2020\|EPI_ISL_744087\|2020 | .................... | .................... | .................... | .................... | .................... | .................... |
| Z | 4315 | hCoV-19/Wales/PHWC-4A334E/2020\|EPI_ISL_744090\|2020 | .................... | .................... | .................... | .................... | .................... | .................... |
| Z | 4317 | hCoV-19/Wales/PHWC-4A33A8/2020\|EPI_ISL_744093\|2020 | .................... | .................... | .................... | .................... | .................... | .................... |
| Z | 4319 | hCoV-19/Wales/PHWC-4A33C6/2020\|EPI_ISL_744095\|2020 | .................... | .................... | .................... | .................... | .................... | .................... |
| Z | 4320 | hCoV-19/Wales/PHWC-4A33D5/2020\|EPI_ISL_744096\|2020 | .................... | .................... | .................... | .................... | .................... | .................... |
| Z | 4321 | hCoV-19/France/ARA-115793/2020\|EPI_ISL_745360\|2020 | .................... | .................... | .................... | .................... | .................... | .................... |
| Z | 4322 | hCoV-19/France/ARA-115811/2020\|EPI_ISL_745361\|2020 | .................... | .................... | .................... | .................... | .................... | .................... |
| Z | 4323 | hCoV-19/France/ARA-115807/2020\|EPI_ISL_745362\|2020 | .................... | .................... | .................... | .................... | .................... | .................... |
| Z | 4324 | hCoV-19/France/ARA-116188/2020\|EPI_ISL_745363\|2020 | .................... | .................... | .................... | .................... | .................... | .................... |
| Z | 4325 | hCoV-19/France/ARA-116443/2020\|EPI_ISL_745364\|2020 | .................... | .................... | .................... | .................... | .................... | .................... |
| Z | 4326 | hCoV-19/France/ARA-116523/2020\|EPI_ISL_745365\|2020 | .................... | .................... | .................... | .................... | .................... | .................... |
| Z | 4327 | hCoV-19/France/ARA-116526/2020\|EPI_ISL_745366\|2020 | .................... | .................... | .................... | .................... | .................... | .................... |
| Z | 4328 | hCoV-19/France/ARA-116531/2020\|EPI_ISL_745367\|2020 | .................... | .................... | .................... | .................... | .................... | .................... |
| Z | 4329 | hCoV-19/France/ARA-116668/2020\|EPI_ISL_745368\|2020 | .................... | .................... | .................... | .................... | .................... | .................... |
| Z | 4330 | hCoV-19/France/ARA-116701/2020\|EPI_ISL_745369\|2020 | .................... | .................... | .................... | .................... | .................... | .................... |
| Z | 4331 | hCoV-19/France/ARA-116737/2020\|EPI_ISL_745370\|2020 | .................... | .................... | .................... | .................... | .................... | .................... |
| Z | 4332 | hCoV-19/France/ARA-117276/2020\|EPI_ISL_745371\|2020 | .................... | .................... | .................... | .................... | .................... | .................... |
| Z | 4333 | hCoV-19/France/ARA-117286/2020\|EPI_ISL_745372\|2020 | .................... | .................... | .................... | .................... | .................... | .................... |
| Z | 4334 | hCoV-19/France/ARA-117399/2020\|EPI_ISL_745373\|2020 | .................... | .................... | .................... | .................... | .................... | .................... |
| Z | 4335 | hCoV-19/France/ARA-117546/2020\|EPI_ISL_745374\|2020 | .................... | .................... | .................... | .................... | .................... | .................... |
| Z | 4336 | hCoV-19/France/ARA-117740/2020\|EPI_ISL_745375\|2020 | .................... | .................... | .................... | .................... | .................... | .................... |
| Z | 4337 | hCoV-19/France/ARA-117764/2020\|EPI_ISL_745376\|2020 | .................... | .................... | .................... | .................... | .................... | .................... |
| Z | 4338 | hCoV-19/France/ARA-117777/2020\|EPI_ISL_745377\|2020 | .................... | .................... | .................... | .................... | .................... | .................... |
| Z | 4339 | hCoV-19/France/ARA-118441/2020\|EPI_ISL_745378\|2020 | .................... | .................... | .................... | .................... | .................... | .................... |
| Z | 4340 | hCoV-19/France/ARA-118640/2020\|EPI_ISL_745379\|2020 | .................... | .................... | .................... | .................... | .................... | .................... |
| Z | 4341 | hCoV-19/France/ARA-119031/2020\|EPI_ISL_745383\|2020 | .................... | .................... | .................... | .................... | .................... | .................... |
| Z | 4342 | hCoV-19/France/ARA-119137/2020\|EPI_ISL_745384\|2020 | .................... | .................... | .................... | .................... | .................... | .................... |
| Z | 4343 | hCoV-19/Chile/RM-164001/2020\|EPI_ISL_746690\|2020-0 | .................... | .................... | .................... | .................... | .................... | .................... |
| Z | 4344 | hCoV-19/Chile/RM-164078/2020\|EPI_ISL_746691\|2020-0 | .................... | .................... | .................... | .................... | .................... | .................... |
| Z | 4345 | hCoV-19/Chile/RM-164508/2020\|EPI_ISL_746692\|2020-0 | .................... | .................... | .................... | .................... | .................... | .................... |
| Z | 4346 | hCoV-19/Chile/RM-164522/2020\|EPI_ISL_746693\|2020-0 | .................... | .................... | .................... | .................... | .................... | .................... |
| Z | 4347 | hCoV-19/Chile/RM-164804/2020\|EPI_ISL_746694\|2020-0 | .................... | .................... | .................... | .................... | .................... | .................... |
| Z | 4348 | hCoV-19/Chile/RM-165835/2020\|EPI_ISL_746695\|2020-0 | .................... | .................... | .................... | .................... | .................... | .................... |
| Z | 4349 | hCoV-19/Chile/RM-165839/2020\|EPI_ISL_746696\|2020-0 | .................... | .................... | .................... | .................... | .................... | .................... |
| Z | 4350 | hCoV-19/Chile/RM-165843/2020\|EPI_ISL_746697\|2020-0 | .................... | .................... | .................... | .................... | .................... | .................... |
| Z | 4351 | hCoV-19/Chile/RM-166412/2020\|EPI_ISL_746698\|2020-0 | .................... | .................... | .................... | .................... | .................... | .................... |
| Z | 4352 | hCoV-19/Chile/RM-166624/2020\|EPI_ISL_746699\|2020-0 | .................... | .................... | .................... | .................... | .................... | .................... |
| Z | 4353 | hCoV-19/USA/CA-CZB-15056/2020\|EPI_ISL_739560\|2020- | .................... | .A.................. | .................... | .................... | .................... | .................... |
| Z | 4354 | hCoV-19/USA/CA-CZB-14516/2020\|EPI_ISL_739561\|2020- | .................... | .................... | .................... | .................... | .................... | .................... |

| | | | | | | | |
|---|---|---|---|---|---|---|---|
| Z | 4355 hCoV-19/USA/CA-CZB-15315/2020\|EPI_ISL_739562\|2020- | ···················· | ·A·················· | ···················· | ···················· | ···················· | ···················· |
| Z | 4356 hCoV-19/USA/CA-CZB-14298/2020\|EPI_ISL_739563\|2020- | ···················· | ·A·················· | ···················· | ···················· | ···················· | ···················· |
| Z | 4357 hCoV-19/USA/CA-CZB-16050/2020\|EPI_ISL_739564\|2020- | ···················· | ···················· | ···················· | ···················· | ···················· | ···················· |
| Z | 4358 hCoV-19/USA/CA-CZB-14260/2020\|EPI_ISL_739565\|2020- | ···················· | ···················· | ···················· | ···················· | ···················· | ···················· |
| Z | 4359 hCoV-19/USA/CA-CZB-15402/2020\|EPI_ISL_739566\|2020- | ···················· | ···················· | ···················· | ···················· | ···················· | ···················· |
| Z | 4361 hCoV-19/USA/CA-CZB-14643/2020\|EPI_ISL_739568\|2020- | ···················· | ···················· | ···················· | ···················· | ···················· | ···················· |
| Z | 4362 hCoV-19/USA/CA-CZB-13956/2020\|EPI_ISL_739569\|2020- | ···················· | ···················· | ···················· | ···················· | ···················· | ···················· |
| Z | 4363 hCoV-19/USA/CA-CZB-14707/2020\|EPI_ISL_739570\|2020- | ···················· | ···················· | ···················· | ···················· | ···················· | ···················· |
| Z | 4364 hCoV-19/USA/CA-CZB-14729/2020\|EPI_ISL_739571\|2020- | ···················· | ·A·················· | ···········T········ | ···················· | ···················· | ···················· |
| Z | 4365 hCoV-19/USA/CA-CZB-13922/2020\|EPI_ISL_739572\|2020- | ···················· | ·A·················· | ···················· | ···················· | ···················· | ···················· |
| Z | 4366 hCoV-19/USA/CA-CZB-14664/2020\|EPI_ISL_739573\|2020- | ···················· | ···················· | ···················· | ···················· | ···················· | ···················· |
| Z | 4367 hCoV-19/USA/CA-CZB-15060/2020\|EPI_ISL_739574\|2020- | ···················· | ···················· | ···················· | ···················· | ···················· | ···················· |
| Z | 4370 hCoV-19/USA/CA-CZB-14938/2020\|EPI_ISL_739577\|2020- | ···················· | ·A·················· | ···················· | ···················· | ···················· | ···················· |
| Z | 4372 hCoV-19/USA/CA-CZB-15318/2020\|EPI_ISL_739579\|2020- | ···················· | ·A·················· | ···················· | ···················· | ···················· | ···················· |
| Z | 4373 hCoV-19/USA/CA-CZB-13669/2020\|EPI_ISL_739580\|2020- | ···················· | ···················· | ···················· | ···················· | ···················· | ···················· |
| Z | 4374 hCoV-19/USA/CA-CZB-14880/2020\|EPI_ISL_739581\|2020- | ···················· | ···················· | ···················· | ···················· | ···················· | ···················· |
| Z | 4375 hCoV-19/USA/CA-CZB-14638/2020\|EPI_ISL_739582\|2020- | ···················· | ···················· | ···················· | ···················· | ···················· | ···················· |
| Z | 4377 hCoV-19/USA/CA-CZB-15224/2020\|EPI_ISL_739584\|2020- | ···················· | ···················· | ···················· | ···················· | ···················· | ···················· |
| Z | 4378 hCoV-19/USA/CA-CZB-14844/2020\|EPI_ISL_739585\|2020- | ···················· | ···················· | ···················· | ···················· | ···················· | ···················· |
| Z | 4379 hCoV-19/USA/CA-CZB-15465/2020\|EPI_ISL_739586\|2020- | ···················· | ···················· | ···T················ | ···················· | ···················· | ···················· |
| Z | 4380 hCoV-19/USA/CA-CZB-14056/2020\|EPI_ISL_739587\|2020- | ···················· | ···················· | ···················· | ···················· | ···················· | ···················· |
| Z | 4381 hCoV-19/USA/CA-CZB-15488/2020\|EPI_ISL_739588\|2020- | ···················· | ·A·················· | ···················· | ···················· | ···················· | ···················· |
| Z | 4383 hCoV-19/USA/CA-CZB-15983/2020\|EPI_ISL_739593\|2020- | ···················· | ·A·················· | ···················· | ···················· | ···················· | ···················· |
| Z | 4384 hCoV-19/USA/CA-CZB-15124/2020\|EPI_ISL_739594\|2020- | ···················· | ·A·················· | ···················· | ···················· | ···················· | ···················· |
| Z | 4385 hCoV-19/USA/CA-CZB-13720/2020\|EPI_ISL_739595\|2020- | ···················· | ···················· | ···················· | ···················· | ···················· | ···················· |
| Z | 4386 hCoV-19/USA/CA-CZB-14975/2020\|EPI_ISL_739596\|2020- | ···················· | ···················· | ···················· | ···················· | ···················· | ···················· |
| Z | 4387 hCoV-19/USA/CA-CZB-15183/2020\|EPI_ISL_739597\|2020- | ···················· | ·A·················· | ···················· | ···················· | ···················· | ···················· |
| Z | 4388 hCoV-19/USA/CA-CZB-14344/2020\|EPI_ISL_739598\|2020- | ···················· | ···················· | ···················· | ···················· | ···················· | ···················· |
| Z | 4389 hCoV-19/Luxembourg/LNS4781293/2020\|EPI_ISL_740544\| | ···················· | ···················· | ···················· | ···················· | ···················· | ···················· |
| Z | 4390 hCoV-19/Luxembourg/LNS4787447/2020\|EPI_ISL_740545\| | ···················· | ···················· | ···················· | ···················· | ···················· | ···················· |
| Z | 4391 hCoV-19/Luxembourg/LNS4788360/2020\|EPI_ISL_740546\| | ···················· | ···················· | ···················· | ···················· | ···················· | ···················· |
| Z | 4392 hCoV-19/Taiwan/NTU37/2020\|EPI_ISL_740547\|2020-11-2 | ···················· | ···················· | ···················· | ···················· | ···················· | ···················· |
| Z | 4393 hCoV-19/Canada/BC-BCCDC-6390/2020\|EPI_ISL_740548\|2 | ···················· | ···················· | ···················· | ···················· | ···················· | ···················· |
| Z | 4394 hCoV-19/Canada/BC-BCCDC-6392/2020\|EPI_ISL_740550\|2 | ···················· | ···················· | ···················· | ···················· | ···················· | ···················· |
| Z | 4395 hCoV-19/Canada/BC-BCCDC-6394/2020\|EPI_ISL_740551\|2 | ···················· | ·A·················· | ···················· | ···················· | ···················· | ···A················ |
| Z | 4396 hCoV-19/Canada/BC-BCCDC-6396/2020\|EPI_ISL_740553\|2 | ···················· | ···················· | ···················· | ···················· | ···················· | ···················· |
| Z | 4397 hCoV-19/Canada/BC-BCCDC-6398/2020\|EPI_ISL_740554\|2 | ···················· | ···················· | ···················· | ···················· | ···················· | ···················· |
| Z | 4398 hCoV-19/Canada/BC-BCCDC-6399/2020\|EPI_ISL_740555\|2 | ···················· | ···················· | ···················· | ···················· | ···················· | ···················· |
| Z | 4399 hCoV-19/Canada/BC-BCCDC-6401/2020\|EPI_ISL_740557\|2 | ···················· | ···················· | ···················· | ···················· | ···················· | ···················· |
| Z | 4400 hCoV-19/Canada/BC-BCCDC-6404/2020\|EPI_ISL_740559\|2 | ···················· | ·A·················· | ···················· | ···················· | ···················· | ···A················ |
| Z | 4401 hCoV-19/Canada/BC-BCCDC-6405/2020\|EPI_ISL_740560\|2 | ···················· | ·A·················· | ···················· | ···················· | ···················· | ···A················ |
| Z | 4402 hCoV-19/Canada/BC-BCCDC-6406/2020\|EPI_ISL_740561\|2 | ···················· | ·A·················· | ···················· | ···················· | ···················· | ···A················ |
| Z | 4403 hCoV-19/Canada/BC-BCCDC-6407/2020\|EPI_ISL_740562\|2 | ···················· | ···················· | ···················· | ···················· | ···················· | ···················· |
| Z | 4404 hCoV-19/Canada/BC-BCCDC-6408/2020\|EPI_ISL_740563\|2 | ···················· | ···················· | ···················· | ···················· | ···················· | ···················· |
| Z | 4405 hCoV-19/Canada/BC-BCCDC-6409/2020\|EPI_ISL_740564\|2 | ···················· | ···················· | ···················· | ···················· | ···················· | ···················· |
| Z | 4406 hCoV-19/Canada/BC-BCCDC-6410/2020\|EPI_ISL_740565\|2 | ···················· | ···················· | ···················· | ···················· | ···················· | ···················· |
| Z | 4407 hCoV-19/Canada/BC-BCCDC-6411/2020\|EPI_ISL_740566\|2 | ···················· | ···················· | ···················· | ···················· | ···················· | ···················· |
| Z | 4408 hCoV-19/Canada/BC-BCCDC-6412/2020\|EPI_ISL_740567\|2 | ···················· | ···················· | ···················· | ···················· | ···················· | ···················· |
| Z | 4409 hCoV-19/Canada/BC-BCCDC-6413/2020\|EPI_ISL_740568\|2 | ···················· | ···················· | ···················· | ···················· | ···················· | ···················· |
| Z | 4410 hCoV-19/Canada/BC-BCCDC-6414/2020\|EPI_ISL_740569\|2 | ···················· | ·A·················· | ···················· | ···················· | ···················· | ···A················ |
| Z | 4411 hCoV-19/Canada/BC-BCCDC-6416/2020\|EPI_ISL_740570\|2 | ···················· | ···················· | ···················· | ···················· | ···················· | ···················· |
| Z | 4412 hCoV-19/Canada/BC-BCCDC-6417/2020\|EPI_ISL_740571\|2 | ···················· | ···················· | ···················· | ···················· | ···················· | ···················· |
| Z | 4413 hCoV-19/Canada/BC-BCCDC-6418/2020\|EPI_ISL_740572\|2 | ···················· | ···················· | ···················· | ···················· | ···················· | ···················· |
| Z | 4414 hCoV-19/Canada/BC-BCCDC-6419/2020\|EPI_ISL_740573\|2 | ···················· | ···················· | ···················· | ···················· | ···················· | ···················· |
| Z | 4415 hCoV-19/Canada/BC-BCCDC-6420/2020\|EPI_ISL_740574\|2 | ···················· | ·A·················· | ···················· | ···················· | ···················· | ···A················ |
| Z | 4416 hCoV-19/Canada/BC-BCCDC-6421/2020\|EPI_ISL_740575\|2 | ···················· | ·A·················· | ···················· | ···················· | ···················· | ···A················ |
| Z | 4417 hCoV-19/Canada/BC-BCCDC-6422/2020\|EPI_ISL_740576\|2 | ···················· | ·A·················· | ···················· | ···················· | ···················· | ···A················ |
| Z | 4418 hCoV-19/Canada/BC-BCCDC-6426/2020\|EPI_ISL_740579\|2 | ···················· | ·A·················· | ···················· | ···················· | ···················· | ···A················ |
| Z | 4419 hCoV-19/Canada/BC-BCCDC-6436/2020\|EPI_ISL_740588\|2 | ···················· | ···················· | ···················· | ···················· | ···················· | ···················· |
| Z | 4420 hCoV-19/Canada/BC-BCCDC-6437/2020\|EPI_ISL_740589\|2 | ···················· | ···················· | ···················· | ···················· | ···················· | ···················· |
| Z | 4421 hCoV-19/Canada/BC-BCCDC-6438/2020\|EPI_ISL_740590\|2 | ···················· | ···················· | ···················· | ···················· | ···················· | ···················· |
| Z | 4422 hCoV-19/Canada/BC-BCCDC-6439/2020\|EPI_ISL_740591\|2 | ···················· | ·A·················· | ···················· | ···················· | ···················· | ···A················ |
| Z | 4423 hCoV-19/Canada/BC-BCCDC-6440/2020\|EPI_ISL_740592\|2 | ···················· | ·A·················· | ···················· | ···················· | ···················· | ···A················ |
| Z | 4424 hCoV-19/Canada/BC-BCCDC-6445/2020\|EPI_ISL_740595\|2 | ···················· | ···················· | ···················· | ···················· | ···················· | ···················· |
| Z | 4425 hCoV-19/Canada/BC-BCCDC-6446/2020\|EPI_ISL_740596\|2 | ···················· | ···················· | ···················· | ···················· | ···················· | ···················· |
| Z | 4426 hCoV-19/Canada/BC-BCCDC-6449/2020\|EPI_ISL_740597\|2 | ···················· | ·A·················· | ···················· | ···················· | ···················· | ···A················ |
| Z | 4427 hCoV-19/Canada/BC-BCCDC-6450/2020\|EPI_ISL_740598\|2 | ···················· | ···················· | ···················· | ···················· | ···················· | ···················· |
| Z | 4428 hCoV-19/Canada/BC-BCCDC-6451/2020\|EPI_ISL_740599\|2 | ···················· | ···················· | ···················· | ···················· | ···················· | ···················· |
| Z | 4429 hCoV-19/England/QEUH-B7D5D7/2020\|EPI_ISL_741875\|20 | ···················· | ···················· | ···················· | ···················· | ···················· | ···················· |
| Z | 4430 hCoV-19/England/QEUH-B7D5E6/2020\|EPI_ISL_741876\|20 | ···················· | ···················· | ···················· | ···················· | ···················· | ···················· |
| Z | 4431 hCoV-19/England/QEUH-B7D601/2020\|EPI_ISL_741877\|20 | ········T··········· | ···················· | ···················· | ···················· | ···················· | ···················· |
| Z | 4432 hCoV-19/England/QEUH-B7D768/2020\|EPI_ISL_741878\|20 | ···················· | ···················· | ···················· | ···················· | ···················· | ···················· |
| Z | 4433 hCoV-19/England/QEUH-B7D786/2020\|EPI_ISL_741879\|20 | ···················· | ···················· | ···················· | ···················· | ···················· | ···················· |
| Z | 4434 hCoV-19/England/QEUH-B7D7D1/2020\|EPI_ISL_741880\|20 | ···················· | ···················· | ···················· | ···················· | ···················· | ···················· |
| Z | 4435 hCoV-19/England/QEUH-B7D7FF/2020\|EPI_ISL_741881\|20 | ···················· | ···················· | ···················· | ···················· | ···················· | ···················· |
| Z | 4436 hCoV-19/England/QEUH-B7D81A/2020\|EPI_ISL_741882\|20 | ···················· | ···················· | ···················· | ···················· | ···················· | ···················· |
| Z | 4437 hCoV-19/England/QEUH-B7D829/2020\|EPI_ISL_741883\|20 | ···················· | ···················· | ···················· | ···················· | ···················· | ···················· |
| Z | 4438 hCoV-19/England/QEUH-B7D847/2020\|EPI_ISL_741884\|20 | ···················· | ···················· | ···················· | ···················· | ···················· | ···················· |
| Z | 4439 hCoV-19/England/QEUH-B7D856/2020\|EPI_ISL_741885\|20 | ···················· | ···················· | ···················· | ···················· | ···················· | ···················· |
| Z | 4440 hCoV-19/England/QEUH-B7D865/2020\|EPI_ISL_741886\|20 | ···················· | ···················· | ···················· | ···················· | ···················· | ···················· |

| | | | | | | | | |
|---|---|---|---|---|---|---|---|---|
| Z | 4441 hCoV-19/England/QEUH-B7D883/2020\|EPI_ISL_741887\|20 | ···················· | ···················· | ····················· | ···················· | ···················· | ···················· | |
| Z | 4442 hCoV-19/England/QEUH-B7D892/2020\|EPI_ISL_741888\|20 | ···················· | ···················· | ····················· | ···················· | ···················· | ···················· | |
| Z | 4443 hCoV-19/England/QEUH-B7DA7E/2020\|EPI_ISL_741889\|20 | ···················· | ···················· | ····················· | ···················· | ···················· | ···················· | |
| Z | 4444 hCoV-19/England/QEUH-B7DA8D/2020\|EPI_ISL_741890\|20 | ···················· | ···················· | ····················· | ···················· | ···················· | ···················· | |
| Z | 4445 hCoV-19/England/QEUH-B7DA9C/2020\|EPI_ISL_741891\|20 | ···················· | ···················· | ····················· | ···················· | ···················· | ···················· | |
| Z | 4446 hCoV-19/England/QEUH-B7DAAB/2020\|EPI_ISL_741892\|20 | ···················· | ···················· | ····················· | ···················· | ···················· | ···················· | |
| Z | 4447 hCoV-19/England/QEUH-B7DABA/2020\|EPI_ISL_741893\|20 | ···················· | ···················· | ····················· | ···················· | ···················· | ···················· | |
| Z | 4448 hCoV-19/England/QEUH-B7DAD8/2020\|EPI_ISL_741894\|20 | ···················· | ···················· | ····················· | ···················· | ···················· | ···················· | |
| Z | 4449 hCoV-19/England/QEUH-B7DAE7/2020\|EPI_ISL_741895\|20 | ···················· | ···················· | ····················· | ···················· | ···················· | ···················· | |
| Z | 4450 hCoV-19/England/QEUH-B7DAF6/2020\|EPI_ISL_741896\|20 | ···················· | ···················· | ····················· | ···················· | ···················· | ···················· | |
| Z | 4451 hCoV-19/England/QEUH-B7DB02/2020\|EPI_ISL_741897\|20 | ···················· | ···················· | ····················· | ···················· | ···················· | ···················· | |
| Z | 4452 hCoV-19/England/QEUH-B7DB4E/2020\|EPI_ISL_741899\|20 | ···················· | ···················· | ····················· | ···················· | ···················· | ···················· | |
| Z | 4453 hCoV-19/Wales/PHWC-4A344B/2020\|EPI_ISL_744101\|2020 | ···················· | ···················· | ····················· | ···················· | ···················· | ···················· | |
| Z | 4455 hCoV-19/Wales/PHWC-4A34F0/2020\|EPI_ISL_744104\|2020 | ···················· | ···················· | ····················· | ···················· | NNN················· | ···················· | 1 |
| Z | 4457 hCoV-19/Wales/PHWC-4A351B/2020\|EPI_ISL_744106\|2020 | ···················· | ···················· | ····················· | ···················· | ···················· | ···················· | |
| Z | 4459 hCoV-19/Wales/PHWC-4A3672/2020\|EPI_ISL_744122\|2020 | ···················· | ···················· | ····················· | ···················· | ···················· | ···················· | |
| Z | 4460 hCoV-19/Wales/PHWC-4A3681/2020\|EPI_ISL_744123\|2020 | ···················· | ···················· | ····················· | ···················· | ···················· | ···················· | |
| Z | 4461 hCoV-19/Wales/PHWC-4A3690/2020\|EPI_ISL_744124\|2020 | ···················· | ···················· | ····················· | ···················· | ···················· | ···················· | |
| Z | 4462 hCoV-19/Wales/PHWC-4A36BE/2020\|EPI_ISL_744125\|2020 | ···················· | ···················· | ····················· | ···················· | ···················· | ···················· | |
| Z | 4463 hCoV-19/Wales/PHWC-4A36FA/2020\|EPI_ISL_744128\|2020 | ···················· | ···················· | ····················· | ···················· | ···················· | ···················· | |
| Z | 4464 hCoV-19/Wales/PHWC-4A3733/2020\|EPI_ISL_744130\|2020 | ···················· | ···················· | ····················· | ···················· | ···················· | ···················· | |
| Z | 4465 hCoV-19/Wales/PHWC-4A3751/2020\|EPI_ISL_744132\|2020 | ···················· | ···················· | ····················· | ···················· | ···················· | ···················· | |
| Z | 4466 hCoV-19/Wales/PHWC-4A3760/2020\|EPI_ISL_744133\|2020 | ···················· | ···················· | ····················· | ···················· | ···················· | ···················· | |
| Z | 4467 hCoV-19/Wales/PHWC-4A377F/2020\|EPI_ISL_744134\|2020 | ···················· | ···················· | ····················· | ···················· | ···················· | ···················· | |
| Z | 4468 hCoV-19/Luxembourg/LNS4792884/2020\|EPI_ISL_744135\| | ···················· | ···················· | ····················· | ···················· | ···················· | ···················· | |
| Z | 4469 hCoV-19/Luxembourg/LNS4814221/2020\|EPI_ISL_744137\| | ···················· | ···················· | ····················· | ···················· | ···················· | ···················· | |
| Z | 4470 hCoV-19/Luxembourg/LNS4820229/2020\|EPI_ISL_744140\| | ···················· | ···················· | ····················· | ···················· | ···················· | ···················· | |
| Z | 4471 hCoV-19/Luxembourg/LNS4826357/2020\|EPI_ISL_744141\| | ···················· | ···················· | ····················· | ···················· | ···················· | ···················· | |
| Z | 4472 hCoV-19/Luxembourg/LNS4826464/2020\|EPI_ISL_744142\| | ···················· | ···················· | ····················· | ···················· | ···················· | ···················· | |
| Z | 4473 hCoV-19/Luxembourg/LNS4834565/2020\|EPI_ISL_744143\| | ···················· | ···················· | ····················· | ···················· | ···················· | ···················· | |
| Z | 4474 hCoV-19/Luxembourg/LNS4835406/2020\|EPI_ISL_744144\| | ···················· | ···················· | ····················· | ···················· | ···················· | ···················· | |
| Z | 4475 hCoV-19/Luxembourg/LNS4835608/2020\|EPI_ISL_744145\| | ···················· | ···················· | ····················· | ···················· | ···················· | ···················· | |
| Z | 4476 hCoV-19/Luxembourg/LNS4841241/2020\|EPI_ISL_744146\| | ···················· | ···················· | ····················· | ···················· | ···················· | ···················· | |
| Z | 4477 hCoV-19/Luxembourg/LNS4847366/2020\|EPI_ISL_744147\| | ···················· | ···················· | ····················· | ···················· | ···················· | ···················· | |
| Z | 4478 hCoV-19/Luxembourg/LNS4850685/2020\|EPI_ISL_744148\| | ···················· | ···················· | ····················· | ···················· | ···················· | ···················· | |
| Z | 4479 hCoV-19/Luxembourg/LNS4853800/2020\|EPI_ISL_744149\| | ···················· | ···················· | ····················· | ···················· | ···················· | ···················· | |
| Z | 4480 hCoV-19/USA/NY-NYCPHL-001503/2020\|EPI_ISL_745409\|2 | ···················· | ···················· | ····················· | ···················· | ···················· | ···················· | |
| Z | 4481 hCoV-19/USA/NY-NYCPHL-001553/2020\|EPI_ISL_745410\|2 | ···················· | ···················· | ····················· | ···················· | ···················· | ···················· | |
| Z | 4482 hCoV-19/USA/NY-NYCPHL-001514/2020\|EPI_ISL_745412\|2 | ···················· | ···················· | ····················· | ···················· | ···················· | ···················· | |
| Z | 4483 hCoV-19/USA/NY-NYCPHL-001479/2020\|EPI_ISL_745413\|2 | ···················· | ···················· | ····················· | ···················· | ···················· | ···················· | |
| Z | 4484 hCoV-19/USA/NY-NYCPHL-001480/2020\|EPI_ISL_745414\|2 | ···················· | ·A·················· | ····················· | ···················· | ···················· | ···················· | |
| Z | 4485 hCoV-19/USA/NY-NYCPHL-001483/2020\|EPI_ISL_745415\|2 | ···················· | ···················· | ····················· | ···················· | ···················· | ···················· | |
| Z | 4486 hCoV-19/USA/NY-NYCPHL-001484/2020\|EPI_ISL_745416\|2 | ···················· | ···················· | ····················· | ···················· | ···················· | ···················· | |
| Z | 4487 hCoV-19/USA/NY-NYCPHL-001486/2020\|EPI_ISL_745417\|2 | ···················· | ·A·················· | ····················· | ···················· | ···················· | ···················· | |
| Z | 4488 hCoV-19/USA/NY-NYCPHL-001488/2020\|EPI_ISL_745418\|2 | ···················· | ···················· | ····················· | ···················· | ···················· | ···················· | |
| Z | 4489 hCoV-19/USA/NY-NYCPHL-001490/2020\|EPI_ISL_745419\|2 | ···················· | ···················· | ····················· | ···················· | ···················· | ···················· | |
| Z | 4490 hCoV-19/USA/NY-NYCPHL-001491/2020\|EPI_ISL_745420\|2 | ···················· | ·A·················· | ····················· | ···················· | ···················· | ···················· | |
| Z | 4491 hCoV-19/USA/NY-NYCPHL-001493/2020\|EPI_ISL_745421\|2 | ···················· | ···················· | ····················· | ···················· | ···················· | ···················· | |
| Z | 4492 hCoV-19/USA/NY-NYCPHL-001494/2020\|EPI_ISL_745422\|2 | ···················· | ···················· | ····················· | ···················· | ···················· | ···················· | |
| Z | 4493 hCoV-19/USA/NY-NYCPHL-001496/2020\|EPI_ISL_745423\|2 | ···················· | ··················C· | ····················· | ···················· | ···················· | ···················· | 1 |
| Z | 4494 hCoV-19/USA/NY-NYCPHL-001498/2020\|EPI_ISL_745424\|2 | ···················· | ···················· | ····················· | ···················· | ···················· | ···················· | |
| Z | 4495 hCoV-19/USA/NY-NYCPHL-001499/2020\|EPI_ISL_745425\|2 | ···················· | ···················· | ····················· | ···················· | ···················· | ···················· | |
| Z | 4496 hCoV-19/USA/NY-NYCPHL-001501/2020\|EPI_ISL_745426\|2 | ···················· | ···················· | ····················· | ···················· | ···················· | ···················· | |
| Z | 4497 hCoV-19/USA/NY-NYCPHL-001502/2020\|EPI_ISL_745427\|2 | ···················· | ···················· | ····················· | ···················· | ···················· | ···················· | |
| Z | 4498 hCoV-19/USA/NY-NYCPHL-001507/2020\|EPI_ISL_745428\|2 | ···················· | ·A·················· | ····················· | ···················· | ···················· | ···················· | |
| Z | 4499 hCoV-19/USA/NY-NYCPHL-001509/2020\|EPI_ISL_745429\|2 | ···················· | ···················· | ····················· | ···················· | ···················· | ···················· | |
| Z | 4500 hCoV-19/USA/NY-NYCPHL-001511/2020\|EPI_ISL_745430\|2 | ···················· | ···················· | ····················· | ···················· | ···················· | ···················· | |
| Z | 4501 hCoV-19/USA/NY-NYCPHL-001512/2020\|EPI_ISL_745431\|2 | ···················· | ···················· | ····················· | ···················· | ···················· | ···················· | |
| Z | 4502 hCoV-19/USA/NY-NYCPHL-001515/2020\|EPI_ISL_745432\|2 | ···················· | ···················· | ····················· | ···················· | ···················· | ···················· | |
| Z | 4503 hCoV-19/USA/NY-NYCPHL-001516/2020\|EPI_ISL_745433\|2 | ···················· | ···················· | ····················· | ···················· | ···················· | ···················· | |
| Z | 4504 hCoV-19/USA/NY-NYCPHL-001517/2020\|EPI_ISL_745434\|2 | ···················· | ··················C· | ····················· | ···················· | ···················· | ···················· | 1 |
| Z | 4505 hCoV-19/USA/NY-NYCPHL-001518/2020\|EPI_ISL_745435\|2 | ···················· | ···················· | ····················· | ···················· | ···················· | ···················· | |
| Z | 4506 hCoV-19/USA/NY-NYCPHL-001520/2020\|EPI_ISL_745436\|2 | ···················· | ·A·················· | ····················· | ···················· | ···················· | ···················· | |
| Z | 4507 hCoV-19/USA/NY-NYCPHL-001521/2020\|EPI_ISL_745437\|2 | ···················· | ···················· | ···········T········· | ···················· | ···················· | ···················· | |
| Z | 4508 hCoV-19/USA/NY-NYCPHL-001522/2020\|EPI_ISL_745438\|2 | ···················· | ·········A·········· | ····················· | ···················· | ···················· | ···················· | |
| Z | 4509 hCoV-19/USA/NY-NYCPHL-001535/2020\|EPI_ISL_745453\|2 | ···················· | ···················· | ····················· | ···················· | ···················· | ···················· | |
| Z | 4510 hCoV-19/USA/NY-NYCPHL-001536/2020\|EPI_ISL_745454\|2 | ···················· | ···················· | ····················· | ···················· | ···················· | ···················· | |
| Z | 4511 hCoV-19/USA/NY-NYCPHL-001537/2020\|EPI_ISL_745455\|2 | ···················· | ···················· | ····················· | ···················· | ···················· | ···················· | |
| Z | 4512 hCoV-19/USA/NY-NYCPHL-001539/2020\|EPI_ISL_745456\|2 | ···················· | ·············A······ | ····················· | ···················· | ···················· | ···················· | |
| Z | 4513 hCoV-19/USA/NY-NYCPHL-001543/2020\|EPI_ISL_745459\|2 | ···················· | ···················· | ····················· | ···················· | ···················· | ···················· | |
| Z | 4514 hCoV-19/USA/NY-NYCPHL-001544/2020\|EPI_ISL_745460\|2 | ···················· | ···················· | ····················· | ···················· | ···················· | ···················· | |
| Z | 4515 hCoV-19/USA/NY-NYCPHL-001546/2020\|EPI_ISL_745461\|2 | ···················· | ···················· | ····················· | ···················· | ···················· | ···················· | |
| Z | 4516 hCoV-19/USA/NY-NYCPHL-001547/2020\|EPI_ISL_745462\|2 | ···················· | ···················· | ····················· | ···················· | ···················· | ···················· | |
| Z | 4517 hCoV-19/USA/NY-NYCPHL-001548/2020\|EPI_ISL_745463\|2 | ···················· | ···················· | ····················· | ···················· | ···················· | ···················· | |
| Z | 4518 hCoV-19/USA/NY-NYCPHL-001550/2020\|EPI_ISL_745464\|2 | ···················· | ·A·················· | ····················· | ···················· | ···················· | ···················· | |
| Z | 4519 hCoV-19/USA/NY-NYCPHL-001555/2020\|EPI_ISL_745465\|2 | ···················· | ···················· | ····················· | ···················· | ···················· | ···················· | |
| Z | 4520 hCoV-19/USA/NY-NYCPHL-001556/2020\|EPI_ISL_745466\|2 | ···················· | ····T··············· | ····················· | ···················· | ···················· | ···················· | |
| Z | 4521 hCoV-19/USA/NY-NYCPHL-001558/2020\|EPI_ISL_745467\|2 | ···················· | ···················· | ····················· | ···················· | ···················· | ···················· | |
| Z | 4522 hCoV-19/USA/NY-NYCPHL-001559/2020\|EPI_ISL_745468\|2 | ···················· | ···················· | ····················· | ···················· | ···················· | ···················· | |
| Z | 4523 hCoV-19/USA/NY-NYCPHL-001560/2020\|EPI_ISL_745469\|2 | ···················· | ·A·················· | ····················· | ···················· | ···················· | ···················· | |

| | | | | | | | | |
|---|---|---|---|---|---|---|---|---|
| Z | 4524 | hCoV-19/USA/NY-NYCPHL-001561/2020\|EPI_ISL_745470\|2 | .................... | .................... | ..................... | .................... | .................... | .................... |
| Z | 4525 | hCoV-19/Chile/RM-167571/2020\|EPI_ISL_746700\|2020-0 | .................... | .................... | ..................... | .................... | .................... | .................... |
| Z | 4526 | hCoV-19/Chile/RM-167590/2020\|EPI_ISL_746701\|2020-0 | .................... | .................... | ..................... | .................... | .................... | .................... |
| Z | 4527 | hCoV-19/Chile/RM-168269/2020\|EPI_ISL_746702\|2020-0 | .................... | .................... | ..................... | .................... | .................... | .................... |
| Z | 4528 | hCoV-19/Chile/RM-168869/2020\|EPI_ISL_746703\|2020-0 | .................... | .................... | ..................... | .................... | .................... | .................... |
| Z | 4529 | hCoV-19/Chile/RM-168883/2020\|EPI_ISL_746704\|2020-0 | .................... | .................... | ..................... | .................... | .................... | .................... |
| Z | 4530 | hCoV-19/Chile/RM-168934/2020\|EPI_ISL_746705\|2020-0 | .................... | .................... | ..................... | .................... | .................... | .................... |
| Z | 4531 | hCoV-19/Chile/RM-169155/2020\|EPI_ISL_746706\|2020-0 | .................... | .................... | ..................... | .................... | .................... | .................... |
| Z | 4532 | hCoV-19/Chile/RM-169384/2020\|EPI_ISL_746707\|2020-0 | .................... | .................... | ..................... | .................... | .................... | .................... |
| Z | 4533 | hCoV-19/Chile/RM-170031/2020\|EPI_ISL_746708\|2020-0 | .................... | .................... | ..................... | .................... | .................... | .................... |
| Z | 4534 | hCoV-19/Chile/RM-170043/2020\|EPI_ISL_746709\|2020-0 | .................... | .................... | ..................... | .................... | .................... | .................... |
| Z | 4535 | hCoV-19/Chile/RM-170122/2020\|EPI_ISL_746710\|2020-0 | .................... | .................... | ..................... | .................... | .................... | .................... |
| Z | 4536 | hCoV-19/Chile/RM-170137/2020\|EPI_ISL_746711\|2020-0 | .................... | .................... | ..................... | .................... | .................... | .................... |
| Z | 4537 | hCoV-19/Chile/RM-170148/2020\|EPI_ISL_746712\|2020-0 | .................... | .................... | ..................... | .................... | .................... | .................... |
| Z | 4538 | hCoV-19/Chile/RM-170158/2020\|EPI_ISL_746713\|2020-0 | .................... | .................... | ..................... | .................... | .................... | .................... |
| Z | 4539 | hCoV-19/Chile/RM-170315/2020\|EPI_ISL_746714\|2020-0 | .................... | .................... | ..................... | .................... | .................... | .................... |
| Z | 4540 | hCoV-19/Chile/RM-170366/2020\|EPI_ISL_746715\|2020-0 | .................... | .................... | ..................... | .................... | .................... | .................... |
| Z | 4541 | hCoV-19/Chile/RM-170368/2020\|EPI_ISL_746716\|2020-0 | .................... | .................... | ..................... | .................... | .................... | .................... |
| Z | 4542 | hCoV-19/Chile/RM-170370/2020\|EPI_ISL_746717\|2020-0 | .................... | .................... | ..................... | .................... | .................... | .................... |
| Z | 4543 | hCoV-19/Chile/RM-170468/2020\|EPI_ISL_746718\|2020-0 | .................... | .................... | ..................... | .................... | .................... | .................... |
| Z | 4544 | hCoV-19/Chile/RM-170499/2020\|EPI_ISL_746719\|2020-0 | .................... | .................... | ..................... | .................... | .................... | .................... |
| Z | 4545 | hCoV-19/Chile/RM-170589/2020\|EPI_ISL_746720\|2020-0 | .................... | .................... | ..................... | .................... | .................... | .................... |
| Z | 4546 | hCoV-19/Chile/RM-170707/2020\|EPI_ISL_746721\|2020-0 | .................... | .................... | ..................... | .................... | .................... | .................... |
| Z | 4547 | hCoV-19/Chile/RM-170921/2020\|EPI_ISL_746722\|2020-0 | .................... | .................... | ..................... | .................... | .................... | .................... |
| Z | 4548 | hCoV-19/Chile/RM-170979/2020\|EPI_ISL_746723\|2020-0 | .................... | .................... | ..................... | .................... | .................... | .................... |
| Z | 4549 | hCoV-19/Chile/RM-170990/2020\|EPI_ISL_746724\|2020-0 | .................... | .................... | ..................... | .................... | .................... | .................... |
| Z | 4550 | hCoV-19/Chile/RM-171011/2020\|EPI_ISL_746725\|2020-0 | .................... | .................... | ..................... | .................... | .................... | .................... |
| Z | 4551 | hCoV-19/Chile/RM-171015/2020\|EPI_ISL_746726\|2020-0 | .................... | .................... | ..................... | .................... | .................... | .................... |
| Z | 4552 | hCoV-19/Chile/RM-171316/2020\|EPI_ISL_746727\|2020-0 | .................... | .................... | ..................... | .................... | .................... | .................... |
| Z | 4553 | hCoV-19/Chile/RM-171327/2020\|EPI_ISL_746728\|2020-0 | .................... | .................... | ..................... | .................... | .................... | .................... |
| Z | 4554 | hCoV-19/Chile/RM-171329/2020\|EPI_ISL_746729\|2020-0 | .................... | .................... | ..................... | .................... | .................... | .................... |
| Z | 4555 | hCoV-19/Chile/RM-171340/2020\|EPI_ISL_746730\|2020-0 | .................... | .................... | ..................... | .................... | .................... | .................... |
| Z | 4556 | hCoV-19/Chile/RM-183871/2020\|EPI_ISL_746731\|2020-0 | .................... | .................... | ..................... | .................... | .................... | .................... |
| Z | 4557 | hCoV-19/Chile/RM-184415/2020\|EPI_ISL_746732\|2020-0 | .................... | .................... | ..................... | .................... | .................... | .................... |
| Z | 4558 | hCoV-19/Chile/RM-185039/2020\|EPI_ISL_746733\|2020-0 | .................... | .................... | ..................... | .................... | .................... | .................... |
| Z | 4559 | hCoV-19/Chile/RM-185819/2020\|EPI_ISL_746734\|2020-0 | .................... | .................... | ..................... | .................... | .................... | .................... |
| Z | 4560 | hCoV-19/Chile/RM-185867/2020\|EPI_ISL_746735\|2020-0 | .................... | .................... | ..................... | .................... | .................... | .................... |
| Z | 4561 | hCoV-19/Chile/RM-187626/2020\|EPI_ISL_746736\|2020-0 | .................... | .................... | ..................... | .................... | .................... | .................... |
| Z | 4562 | hCoV-19/Chile/RM-187968/2020\|EPI_ISL_746737\|2020-0 | .................... | .................... | ..................... | .................... | .................... | .................... |
| Z | 4563 | hCoV-19/Chile/RM-188811/2020\|EPI_ISL_746738\|2020-0 | .................... | .................... | ..................... | .................... | .................... | .................... |
| Z | 4564 | hCoV-19/Chile/RM-188963/2020\|EPI_ISL_746739\|2020-0 | .................... | .................... | ..................... | .................... | .................... | .................... |
| Z | 4565 | hCoV-19/Chile/RM-189170/2020\|EPI_ISL_746740\|2020-0 | .................... | .................... | ..................... | .................... | .................... | .................... |
| Z | 4566 | hCoV-19/Chile/RM-189585/2020\|EPI_ISL_746741\|2020-0 | .................... | .................... | ..................... | .................... | .................... | .................... |
| Z | 4567 | hCoV-19/Chile/RM-190290/2020\|EPI_ISL_746742\|2020-0 | .................... | .................... | ..................... | .................... | .................... | .................... |
| Z | 4568 | hCoV-19/Chile/RM-190581/2020\|EPI_ISL_746743\|2020-0 | .................... | .................... | ..................... | .................... | .................... | .................... |
| Z | 4569 | hCoV-19/Chile/RM-190882/2020\|EPI_ISL_746744\|2020-0 | .................... | .................... | ..................... | .................... | .................... | .................... |
| Z | 4570 | hCoV-19/Chile/RM-191185/2020\|EPI_ISL_746745\|2020-0 | .................... | .................... | ..................... | .................... | .................... | .................... |
| Z | 4571 | hCoV-19/Chile/RM-191226/2020\|EPI_ISL_746746\|2020-0 | .................... | .................... | ..................... | .................... | .................... | .................... |
| Z | 4572 | hCoV-19/Chile/RM-191308/2020\|EPI_ISL_746747\|2020-0 | .................... | .................... | ..................... | .................... | .................... | .................... |
| Z | 4573 | hCoV-19/Chile/RM-191591/2020\|EPI_ISL_746748\|2020-0 | .................... | .................... | ..................... | .................... | .................... | .................... |
| Z | 4574 | hCoV-19/Chile/RM-191594/2020\|EPI_ISL_746749\|2020-0 | .................... | .................... | ..................... | .................... | .................... | .................... |
| Z | 4575 | hCoV-19/Chile/RM-191841/2020\|EPI_ISL_746750\|2020-0 | .................... | .................... | ..................... | .................... | .................... | .................... |
| Z | 4576 | hCoV-19/Chile/RM-191972/2020\|EPI_ISL_746751\|2020-0 | .................... | .................... | ..................... | .................... | .................... | .................... |
| Z | 4577 | hCoV-19/Chile/RM-192950/2020\|EPI_ISL_746752\|2020-0 | .................... | .................... | ..................... | .................... | .................... | .................... |
| Z | 4578 | hCoV-19/Chile/RM-193052/2020\|EPI_ISL_746753\|2020-0 | .................... | .................... | ..................... | .................... | .................... | .................... |
| Z | 4579 | hCoV-19/Chile/RM-193465/2020\|EPI_ISL_746754\|2020-0 | .................... | .................... | ..................... | .................... | .................... | .................... |
| Z | 4580 | hCoV-19/Chile/RM-193870/2020\|EPI_ISL_746755\|2020-0 | .................... | .................... | ..................... | .................... | .................... | .................... |
| Z | 4581 | hCoV-19/Chile/RM-194896/2020\|EPI_ISL_746756\|2020-0 | .................... | .................... | ..................... | .................... | .................... | .................... |
| Z | 4582 | hCoV-19/Chile/RM-203885/2020\|EPI_ISL_746757\|2020-1 | .................... | .................... | ..................... | .................... | .................... | .................... |
| Z | 4583 | hCoV-19/Chile/RM-203933/2020\|EPI_ISL_746758\|2020-1 | .................... | .................... | ..................... | .................... | .................... | .................... |
| Z | 4584 | hCoV-19/Chile/RM-203937/2020\|EPI_ISL_746759\|2020-1 | .................... | .................... | ..................... | .................... | .................... | .................... |
| Z | 4585 | hCoV-19/Chile/RM-204293/2020\|EPI_ISL_746760\|2020-1 | .................... | .................... | ..................... | .................... | .................... | .................... |
| Z | 4586 | hCoV-19/Chile/RM-204371/2020\|EPI_ISL_746761\|2020-1 | .................... | .................... | ..................... | .................... | .................... | .................... |
| Z | 4587 | hCoV-19/Chile/RM-204484/2020\|EPI_ISL_746762\|2020-1 | .................... | .................... | ..................... | .................... | .................... | .................... |
| Z | 4588 | hCoV-19/Chile/RM-204488/2020\|EPI_ISL_746763\|2020-1 | .................... | .................... | ..................... | .................... | .................... | .................... |
| Z | 4589 | hCoV-19/Chile/RM-204591/2020\|EPI_ISL_746764\|2020-1 | .................... | .................... | ..................... | .................... | .................... | .................... |
| Z | 4590 | hCoV-19/Chile/RM-205625/2020\|EPI_ISL_746765\|2020-1 | .................... | .................... | ..................... | .................... | .................... | .................... |
| Z | 4591 | hCoV-19/Chile/RM-206395/2020\|EPI_ISL_746766\|2020-1 | .................... | .................... | ..................... | .................... | .................... | .................... |
| Z | 4592 | hCoV-19/Chile/RM-207231/2020\|EPI_ISL_746767\|2020-1 | .................... | .................... | ..................... | .................... | .................... | .................... |
| Z | 4593 | hCoV-19/Chile/RM-207382/2020\|EPI_ISL_746768\|2020-1 | .................... | .................... | ..................... | .................... | .................... | .................... |
| Z | 4594 | hCoV-19/Chile/RM-207982/2020\|EPI_ISL_746769\|2020-1 | .................... | .................... | ..................... | .................... | .................... | .................... |
| Z | 4595 | hCoV-19/Chile/RM-72850/2020\|EPI_ISL_746784\|2020-05 | .................... | .................... | ..................... | .................... | .................... | .................... |
| Z | 4596 | hCoV-19/Chile/RM-72851/2020\|EPI_ISL_746785\|2020-05 | .................... | .................... | ..................... | .................... | .................... | .................... |
| Z | 4597 | hCoV-19/Chile/RM-72852/2020\|EPI_ISL_746786\|2020-05 | .................... | .................... | ..................... | .................... | .................... | .................... |
| Z | 4598 | hCoV-19/Chile/RM-73219/2020\|EPI_ISL_746787\|2020-05 | .................... | .................... | ..................... | .................... | .................... | .................... |
| Z | 4599 | hCoV-19/Chile/RM-73254/2020\|EPI_ISL_746790\|2020-05 | .................... | .................... | ..................... | .................... | .................... | .................... |
| Z | 4600 | hCoV-19/Chile/RM-73255/2020\|EPI_ISL_746791\|2020-05 | .................... | .................... | ..................... | .................... | .................... | .................... |
| Z | 4601 | hCoV-19/Chile/RM-82478/2020\|EPI_ISL_746792\|2020-05 | .................... | .................... | ..................... | .................... | .................... | .................... |
| Z | 4602 | hCoV-19/Chile/RM-82488/2020\|EPI_ISL_746793\|2020-05 | .................... | .................... | ..................... | .................... | .................... | .................... |
| Z | 4603 | hCoV-19/Chile/RM-82495/2020\|EPI_ISL_746794\|2020-05 | .................... | .................... | ..................... | .................... | .................... | .................... |

```
Z    4604 hCoV-19/Chile/RM-83539/2020|EPI_ISL_746795|2020-05   ····················  ····················  ····················  ····················  ····················  ····················
Z    4605 hCoV-19/Chile/RM-83540/2020|EPI_ISL_746796|2020-05   ····················  ····················  ····················  ····················  ····················  ····················
Z    4606 hCoV-19/Chile/RM-83548/2020|EPI_ISL_746797|2020-05   ····················  ····················  ····················  ····················  ····················  ····················
Z    4607 hCoV-19/Chile/RM-83553/2020|EPI_ISL_746798|2020-05   ····················  ····················  ····················  ····················  ····················  ····················
Z    4608 hCoV-19/Chile/RM-83568/2020|EPI_ISL_746799|2020-05   ····················  ····················  ····················  ····················  ····················  ····················
Z    4610 hCoV-19/USA/CA-CZB-14127/2020|EPI_ISL_739601|2020-   ····················  ····················  ····················  ····················  ····················  ····················
Z    4611 hCoV-19/USA/CA-CZB-15294/2020|EPI_ISL_739602|2020-   ····················  ·A··················  ····················  ····················  ····················  ····················
Z    4612 hCoV-19/USA/CA-CZB-14629/2020|EPI_ISL_739603|2020-   ····················  ····················  ····················  ····················  ····················  ····················
Z    4613 hCoV-19/USA/CA-CZB-13877/2020|EPI_ISL_739604|2020-   ····················  ·A··················  ····················  ····················  ····················  ····················
Z    4614 hCoV-19/USA/CA-CZB-13689/2020|EPI_ISL_739605|2020-   ····················  ····················  ····················  ····················  ····················  ····················
Z    4615 hCoV-19/USA/CA-CZB-14424/2020|EPI_ISL_739606|2020-   ····················  ····················  ····················  ····················  ····················  ····················
Z    4616 hCoV-19/USA/CA-CZB-14872/2020|EPI_ISL_739607|2020-   ····················  ····················  ····················  ····················  ····················  ····················
Z    4617 hCoV-19/USA/CA-CZB-14860/2020|EPI_ISL_739608|2020-   ····················  ····················  ····················  ····················  ····················  ····················
Z    4619 hCoV-19/USA/CA-CZB-14267/2020|EPI_ISL_739610|2020-   ····················  ·A··················  ····················  ····················  ····················  ····················
Z    4620 hCoV-19/USA/CA-CZB-14804/2020|EPI_ISL_739611|2020-   ····················  ····················  ····················  ····················  ····················  ····················
Z    4621 hCoV-19/USA/CA-CZB-14997/2020|EPI_ISL_739612|2020-   ····················  ····················  ····················  ····················  ····················  ····················
Z    4622 hCoV-19/USA/CA-CZB-14592/2020|EPI_ISL_739613|2020-   ····················  ·A··················  ····················  ····················  ····················  ······R·············
Z    4623 hCoV-19/USA/CA-CZB-14675/2020|EPI_ISL_739614|2020-   ····················  ····················  ····················  ····················  ····················  ····················
Z    4624 hCoV-19/USA/CA-CZB-15351/2020|EPI_ISL_739615|2020-   ····················  ····················  ····················  ····················  ····················  ····················
Z    4626 hCoV-19/USA/CA-CZB-15934/2020|EPI_ISL_739617|2020-   ····················  ····················  ····················  ····················  ····················  ····················
Z    4627 hCoV-19/USA/CA-CZB-13675/2020|EPI_ISL_739618|2020-   ····················  ····················  ····················  ····················  ····················  ····················
Z    4629 hCoV-19/USA/CA-CZB-14203/2020|EPI_ISL_739620|2020-   ····················  ·A··················  ····················  ····················  ····················  ····················
Z    4630 hCoV-19/USA/CA-CZB-14789/2020|EPI_ISL_739621|2020-   ····················  ····················  ····················  ····················  ····················  ····················
Z    4631 hCoV-19/USA/CA-CZB-14020/2020|EPI_ISL_739622|2020-   ····················  ····················  ····················  ····················  ····················  ····················
Z    4633 hCoV-19/USA/CA-CZB-15034/2020|EPI_ISL_739624|2020-   ····················  ·A··················  ····················  ····················  NNN·················  ····················   1
Z    4634 hCoV-19/USA/CA-CZB-14641/2020|EPI_ISL_739625|2020-   ····················  ····················  ····················  ····················  ····················  ····················
Z    4635 hCoV-19/USA/CA-CZB-15169/2020|EPI_ISL_739626|2020-   ····················  ····················  ····················  ····················  ····················  ····················
Z    4636 hCoV-19/USA/CA-CZB-14640/2020|EPI_ISL_739627|2020-   ····················  ····················  ····················  ····················  ····················  ····················
Z    4637 hCoV-19/USA/CA-CZB-14601/2020|EPI_ISL_739628|2020-   ····················  ····················  ····················  ····················  ····················  ····················
Z    4638 hCoV-19/USA/CA-CZB-15176/2020|EPI_ISL_739629|2020-   ····················  ·A··················  ····················  ····················  ····················  ····················
Z    4639 hCoV-19/USA/CA-CZB-16059/2020|EPI_ISL_739630|2020-   ····················  ····················  ····················  ····················  ····················  ····················
Z    4641 hCoV-19/USA/CA-CZB-14594/2020|EPI_ISL_739632|2020-   ····················  ·A··················  ····················  ····················  ····················  ····················
Z    4642 hCoV-19/USA/CA-CZB-14600/2020|EPI_ISL_739633|2020-   ····················  ····················  ····················  ····················  ····················  ····················
Z    4643 hCoV-19/USA/CA-CZB-13759/2020|EPI_ISL_739634|2020-   ····················  ····················  ····················  ····················  ····················  ····················
Z    4644 hCoV-19/USA/CA-CZB-11251/2020|EPI_ISL_739635|2020-   ····················  ····················  ····················  ····················  ····················  ····················
Z    4645 hCoV-19/USA/CA-CZB-14665/2020|EPI_ISL_739636|2020-   ····················  ····················  ····················  ····················  ····················  ····················
Z    4646 hCoV-19/USA/CA-CZB-14216/2020|EPI_ISL_739637|2020-   ····················  ····················  ····················  ····················  ····················  ····················
Z    4647 hCoV-19/USA/CA-CZB-14050/2020|EPI_ISL_739638|2020-   ····················  ···A················  ····················  ····················  ····················  ····················
Z    4648 hCoV-19/USA/CA-CZB-14714/2020|EPI_ISL_739639|2020-   ·······T············  ····················  ····················  ····················  ····················  ····················
Z    4649 hCoV-19/USA/CA-CZB-15464/2020|EPI_ISL_739641|2020-   ····················  ····················  ··T·················  ····················  ····················  ····················
Z    4650 hCoV-19/USA/CA-CZB-15279/2020|EPI_ISL_739642|2020-   ····················  ····················  ····················  ····················  ····················  ·······S············
Z    4651 hCoV-19/USA/CA-CZB-13950/2020|EPI_ISL_739643|2020-   ····················  ····················  ····················  ····················  ····················  ····················
Z    4652 hCoV-19/USA/CA-CZB-13938/2020|EPI_ISL_739644|2020-   ····················  ····················  ····················  ····················  ····················  ····················
Z    4653 hCoV-19/USA/CA-CZB-11276/2020|EPI_ISL_739645|2020-   ····················  ·A··················  ····················  ····················  ····················  ····················
Z    4654 hCoV-19/USA/CA-CZB-14932/2020|EPI_ISL_739646|2020-   ····················  ····················  ····················  ····················  ····················  ····················
Z    4656 hCoV-19/USA/CA-CZB-14378/2020|EPI_ISL_739648|2020-   ····················  ····················  ····················  ····················  ····················  ····················
Z    4657 hCoV-19/USA/CA-CZB-15331/2020|EPI_ISL_739649|2020-   ····················  ·A··················  ····················  ····················  ····················  ····················
Z    4658 hCoV-19/USA/CA-CZB-15957/2020|EPI_ISL_739650|2020-   ····················  ·A··················  ····················  ····················  ····················  ····················
Z    4659 hCoV-19/USA/CA-CZB-14821/2020|EPI_ISL_739651|2020-   ····················  ····················  ····················  ····················  ····················  ····················
Z    4660 hCoV-19/USA/CA-CZB-14101/2020|EPI_ISL_739652|2020-   ····················  ····················  ····················  ····················  ····················  ····················
Z    4661 hCoV-19/USA/CA-CZB-15954/2020|EPI_ISL_739653|2020-   ····················  ····················  ····················  ····················  ····················  ····················
Z    4662 hCoV-19/USA/CA-CZB-14231/2020|EPI_ISL_739654|2020-   ····················  ····················  ····················  ····················  ····················  ····················
Z    4663 hCoV-19/USA/CA-CZB-16173/2020|EPI_ISL_739655|2020-   ····················  ····················  ····················  ····················  ····················  ····················
Z    4664 hCoV-19/USA/CA-CZB-14019/2020|EPI_ISL_739656|2020-   ····················  ····················  ····················  ····················  ····················  ····················
Z    4666 hCoV-19/USA/CA-CZB-14218/2020|EPI_ISL_739658|2020-   ····················  ····················  ····················  ····················  ····················  ····················
Z    4670 hCoV-19/Canada/BC-BCCDC-6452/2020|EPI_ISL_740600|2   ····················  ····················  ····················  ····················  ····················  ····················
Z    4671 hCoV-19/Canada/BC-BCCDC-6453/2020|EPI_ISL_740601|2   ····················  ····················  ····················  ····················  ····················  ····················
Z    4672 hCoV-19/Canada/BC-BCCDC-6454/2020|EPI_ISL_740602|2   ····················  ·A··················  ····················  ····················  ····················  ···A················
Z    4673 hCoV-19/Canada/BC-BCCDC-6455/2020|EPI_ISL_740603|2   ····················  ····················  ····················  ····················  ····················  ····················
Z    4674 hCoV-19/Canada/BC-BCCDC-6458/2020|EPI_ISL_740606|2   ····················  ·A··················  ····················  ····················  ····················  ···A················
Z    4675 hCoV-19/Canada/BC-BCCDC-6463/2020|EPI_ISL_740608|2   ····················  ····················  ····················  ····················  ····················  ····················
Z    4676 hCoV-19/Canada/BC-BCCDC-6464/2020|EPI_ISL_740609|2   ····················  ·A··················  ····················  ····················  ····················  ···A················
Z    4677 hCoV-19/Canada/BC-BCCDC-6466/2020|EPI_ISL_740611|2   ····················  ····················  ····················  ····················  ····················  ····················
Z    4678 hCoV-19/Canada/BC-BCCDC-6467/2020|EPI_ISL_740612|2   ····················  ····················  ····················  ····················  ····················  ····················
Z    4679 hCoV-19/Canada/BC-BCCDC-6469/2020|EPI_ISL_740614|2   ····················  ····················  ····················  ····················  ····················  ····················
Z    4680 hCoV-19/Canada/BC-BCCDC-6470/2020|EPI_ISL_740615|2   ····················  ····················  ····················  ····················  ····················  ····················
Z    4681 hCoV-19/Canada/BC-BCCDC-6471/2020|EPI_ISL_740616|2   ····················  ·A··················  ····················  ····················  ····················  ···A················
Z    4682 hCoV-19/Canada/BC-BCCDC-6472/2020|EPI_ISL_740617|2   ····················  ····················  ····················  ····················  ····················  ····················
Z    4683 hCoV-19/Canada/BC-BCCDC-6473/2020|EPI_ISL_740618|2   ····················  ·A··················  ····················  ····················  ····················  ···A················
Z    4684 hCoV-19/Canada/BC-BCCDC-6474/2020|EPI_ISL_740619|2   ····················  ·A··················  ····················  ····················  ····················  ···A················
Z    4685 hCoV-19/Canada/BC-BCCDC-6479/2020|EPI_ISL_740620|2   ····················  ····················  ····················  ····················  ····················  ····················
Z    4686 hCoV-19/Canada/BC-BCCDC-6481/2020|EPI_ISL_740621|2   ····················  ····················  ····················  ····················  ····················  ····················
Z    4687 hCoV-19/Canada/BC-BCCDC-6482/2020|EPI_ISL_740622|2   ····················  ····················  ····················  ····················  ····················  ····················
Z    4688 hCoV-19/Canada/BC-BCCDC-7230/2020|EPI_ISL_740623|2   ····················  ·A··················  ····················  ····················  ····················  ···A················
Z    4689 hCoV-19/Canada/BC-BCCDC-7233/2020|EPI_ISL_740625|2   ····················  ····················  ····················  ····················  ····················  ····················
Z    4690 hCoV-19/Canada/BC-BCCDC-7238/2020|EPI_ISL_740630|2   ····················  ····················  ····················  ····················  ····················  ····················
Z    4691 hCoV-19/Canada/BC-BCCDC-7239/2020|EPI_ISL_740631|2   ····················  ·A··················  ····················  ····················  ····················  ···A················
Z    4692 hCoV-19/Canada/BC-BCCDC-7241/2020|EPI_ISL_740632|2   ····················  ·A··················  ····················  ····················  ····················  ···A················
Z    4693 hCoV-19/Canada/BC-BCCDC-7242/2020|EPI_ISL_740633|2   ····················  ·A··················  ····················  ····················  ····················  ···A················
Z    4694 hCoV-19/Canada/BC-BCCDC-7243/2020|EPI_ISL_740634|2   ····················  ·A··················  ····················  ····················  ····················  ···A················
```

| | | | | | | | | |
|---|---|---|---|---|---|---|---|---|
| Z | 4695 | hCoV-19/Canada/BC-BCCDC-7245/2020\|EPI_ISL_740635\|2 | ···················· | ·A·················· | ···················· | ···················· | ···················· | ··A················· |
| Z | 4696 | hCoV-19/Canada/BC-BCCDC-7247/2020\|EPI_ISL_740636\|2 | ···················· | ···················· | ···················· | ···················· | ···················· | ···················· |
| Z | 4697 | hCoV-19/Canada/BC-BCCDC-7248/2020\|EPI_ISL_740637\|2 | ···················· | ·A·················· | ···················· | ···················· | ···················· | ··A················· |
| Z | 4698 | hCoV-19/Canada/BC-BCCDC-7249/2020\|EPI_ISL_740638\|2 | ···················· | ···················· | ···················· | ···················· | ···················· | ···················· |
| Z | 4699 | hCoV-19/Canada/BC-BCCDC-7252/2020\|EPI_ISL_740640\|2 | ···················· | ·A·················· | ···················· | ···················· | ···················· | ··A················· |
| Z | 4700 | hCoV-19/Canada/BC-BCCDC-7253/2020\|EPI_ISL_740641\|2 | ···················· | ·A·················· | ···················· | ···················· | ···················· | ··A················· |
| Z | 4701 | hCoV-19/Canada/BC-BCCDC-7258/2020\|EPI_ISL_740643\|2 | ···················· | ·A·················· | ···················· | ···················· | ···················· | ··A················· |
| Z | 4702 | hCoV-19/Canada/BC-BCCDC-7259/2020\|EPI_ISL_740644\|2 | ···················· | ·A·················· | ···················· | ···················· | ···················· | ··A················· |
| Z | 4703 | hCoV-19/Canada/BC-BCCDC-7262/2020\|EPI_ISL_740645\|2 | ···················· | ···················· | ···················· | ···················· | ···················· | ···················· |
| Z | 4704 | hCoV-19/Canada/BC-BCCDC-7265/2020\|EPI_ISL_740647\|2 | ···················· | ···················· | ···················· | ···················· | ···················· | ···················· |
| Z | 4705 | hCoV-19/Canada/BC-BCCDC-7266/2020\|EPI_ISL_740648\|2 | ···················· | ···················· | ···················· | ···················· | ···················· | ···················· |
| Z | 4706 | hCoV-19/Canada/BC-BCCDC-7268/2020\|EPI_ISL_740649\|2 | ···················· | ·A·················· | ···················· | ···················· | ···················· | ··A················· |
| Z | 4707 | hCoV-19/Canada/BC-BCCDC-7270/2020\|EPI_ISL_740651\|2 | ···················· | ·A·················· | ···················· | ···················· | ···················· | ··A················· |
| Z | 4708 | hCoV-19/Canada/BC-BCCDC-7273/2020\|EPI_ISL_740652\|2 | ···················· | ···················· | ···················· | ···················· | ···················· | ···················· |
| Z | 4709 | hCoV-19/Canada/BC-BCCDC-7274/2020\|EPI_ISL_740653\|2 | ···················· | ···················· | ···················· | ···················· | ···················· | ···················· |
| Z | 4710 | hCoV-19/Canada/BC-BCCDC-7280/2020\|EPI_ISL_740654\|2 | ···················· | ·A·················· | ···················· | ···················· | ···················· | ··A················· |
| Z | 4711 | hCoV-19/Canada/BC-BCCDC-7282/2020\|EPI_ISL_740655\|2 | ···················· | ···················· | ···················· | ···················· | ···················· | ···················· |
| Z | 4712 | hCoV-19/Canada/BC-BCCDC-7283/2020\|EPI_ISL_740656\|2 | ···················· | ·A·················· | ···················· | ···················· | ···················· | ··A················· |
| Z | 4713 | hCoV-19/Canada/BC-BCCDC-7286/2020\|EPI_ISL_740657\|2 | ···················· | ···················· | ···················· | ···················· | ···················· | ···················· |
| Z | 4714 | hCoV-19/Canada/BC-BCCDC-7288/2020\|EPI_ISL_740659\|2 | ···················· | ·A·················· | ···················· | ···················· | ···················· | ··A················· |
| Z | 4715 | hCoV-19/Canada/BC-BCCDC-7289/2020\|EPI_ISL_740660\|2 | ···················· | ···················· | ···················· | ···················· | ···················· | ···················· |
| Z | 4716 | hCoV-19/Canada/BC-BCCDC-7290/2020\|EPI_ISL_740661\|2 | ···················· | ···················· | ···················· | ···················· | ···················· | ···················· |
| Z | 4717 | hCoV-19/Canada/BC-BCCDC-7291/2020\|EPI_ISL_740662\|2 | ···················· | ·A·················· | ···················· | ···················· | ···················· | ··A················· |
| Z | 4718 | hCoV-19/Canada/BC-BCCDC-7293/2020\|EPI_ISL_740663\|2 | ···················· | ·A·················· | ···················· | ···················· | ···················· | ··A················· |
| Z | 4719 | hCoV-19/Canada/BC-BCCDC-7294/2020\|EPI_ISL_740664\|2 | ···················· | ·A·················· | ···················· | ···················· | ···················· | ··A················· |
| Z | 4720 | hCoV-19/England/QEUH-B7DB5D/2020\|EPI_ISL_741900\|20 | ···················· | ···················· | ···················· | ···················· | ···················· | ···················· |
| Z | 4721 | hCoV-19/England/QEUH-B7DB6C/2020\|EPI_ISL_741901\|20 | ···················· | ···················· | ···················· | ···················· | ···················· | ···················· |
| Z | 4722 | hCoV-19/England/QEUH-B7DB99/2020\|EPI_ISL_741902\|20 | ···················· | ···················· | ···················· | ···················· | ···················· | ···················· |
| Z | 4723 | hCoV-19/England/QEUH-B7DBA8/2020\|EPI_ISL_741903\|20 | ···················· | ···················· | ···················· | ···················· | ···················· | ···················· |
| Z | 4724 | hCoV-19/England/QEUH-B7DBC6/2020\|EPI_ISL_741904\|20 | ···················· | ···················· | ···················· | ···················· | ···················· | ···················· |
| Z | 4725 | hCoV-19/England/QEUH-B7DBD5/2020\|EPI_ISL_741905\|20 | ···················· | ···················· | ···················· | ···················· | ···················· | ···················· |
| Z | 4726 | hCoV-19/England/QEUH-B7DC0F/2020\|EPI_ISL_741906\|20 | ···················· | ···················· | ···················· | ···················· | ···················· | ···················· |
| Z | 4727 | hCoV-19/England/QEUH-B7DC1E/2020\|EPI_ISL_741907\|20 | ···················· | ···················· | ···················· | ···················· | ···················· | ···················· |
| Z | 4728 | hCoV-19/England/QEUH-B7DC4B/2020\|EPI_ISL_741908\|20 | ···················· | ·A·················· | ···················· | ···················· | ···················· | ···················· |
| Z | 4729 | hCoV-19/England/QEUH-B7DC5A/2020\|EPI_ISL_741909\|20 | ···················· | ···················· | ···················· | ···················· | ···················· | ···················· |
| Z | 4730 | hCoV-19/England/QEUH-B7DC69/2020\|EPI_ISL_741910\|20 | ···················· | ···················· | ···················· | ···················· | ···················· | ···················· |
| Z | 4731 | hCoV-19/England/QEUH-B7DC78/2020\|EPI_ISL_741911\|20 | ···················· | ···················· | ···················· | ···················· | ···················· | ···················· |
| Z | 4732 | hCoV-19/England/QEUH-B7DC87/2020\|EPI_ISL_741912\|20 | ···················· | ···················· | ···················· | ···················· | ···················· | ···················· |
| Z | 4733 | hCoV-19/England/QEUH-B7DCA5/2020\|EPI_ISL_741913\|20 | ···················· | ···················· | ···················· | ···················· | ···················· | ···················· |
| Z | 4734 | hCoV-19/England/QEUH-B7DCB4/2020\|EPI_ISL_741914\|20 | ···················· | ···················· | ···················· | ···················· | ···················· | ···················· |
| Z | 4735 | hCoV-19/England/QEUH-B7DCE1/2020\|EPI_ISL_741915\|20 | ···················· | ···················· | ···················· | ···················· | ···················· | ···················· |
| Z | 4736 | hCoV-19/England/QEUH-B7DCF0/2020\|EPI_ISL_741916\|20 | ···················· | ···················· | ···················· | ···················· | ···················· | ···················· |
| Z | 4737 | hCoV-19/England/QEUH-B7DD1B/2020\|EPI_ISL_741917\|20 | ···················· | ···················· | ···················· | ···················· | ···················· | ···················· |
| Z | 4738 | hCoV-19/England/QEUH-B7DD39/2020\|EPI_ISL_741919\|20 | ···················· | ···················· | ···················· | ···················· | ···················· | ···················· |
| Z | 4739 | hCoV-19/England/QEUH-B7DD48/2020\|EPI_ISL_741920\|20 | ···················· | ·A·················· | ···················· | ···················· | ···················· | ···················· |
| Z | 4740 | hCoV-19/England/QEUH-B7DDDF/2020\|EPI_ISL_741921\|20 | ···················· | ···················· | ···················· | ···················· | ···················· | ···················· |
| Z | 4741 | hCoV-19/England/QEUH-B7DDEE/2020\|EPI_ISL_741922\|20 | ···················· | ···················· | ···················· | ···················· | ···················· | ···················· |
| Z | 4742 | hCoV-19/England/QEUH-B7DDFD/2020\|EPI_ISL_741923\|20 | ···················· | ···················· | ···················· | ···················· | ···················· | ···················· |
| Z | 4743 | hCoV-19/England/QEUH-B7DEAF/2020\|EPI_ISL_741924\|20 | ···················· | ···················· | ···················· | ···················· | ···················· | ···················· |
| Z | 4744 | hCoV-19/England/QEUH-B7DEEB/2020\|EPI_ISL_741925\|20 | ···················· | ···················· | ···················· | ···················· | ···················· | ···················· |
| Z | 4745 | hCoV-19/England/QEUH-B7DEFA/2020\|EPI_ISL_741926\|20 | ···················· | ···················· | ···················· | ···················· | ···················· | ···················· |
| Z | 4746 | hCoV-19/England/QEUH-B7DF06/2020\|EPI_ISL_741927\|20 | ···················· | ···················· | ···················· | ···················· | ···················· | ···················· |
| Z | 4747 | hCoV-19/England/QEUH-B7DF15/2020\|EPI_ISL_741928\|20 | ···················· | ···················· | ···················· | ···················· | ···················· | ···················· |
| Z | 4748 | hCoV-19/England/QEUH-B7DF33/2020\|EPI_ISL_741929\|20 | ···················· | ···················· | ···················· | ···················· | ···················· | ···················· |
| Z | 4749 | hCoV-19/England/QEUH-B7DF60/2020\|EPI_ISL_741930\|20 | ···················· | ···················· | ···················· | ···················· | ···················· | ···················· |
| Z | 4750 | hCoV-19/England/QEUH-B7DF7F/2020\|EPI_ISL_741931\|20 | ···················· | ···················· | ···················· | ···················· | ···················· | ···················· |
| Z | 4751 | hCoV-19/England/QEUH-B7DFCA/2020\|EPI_ISL_741933\|20 | ···················· | ···················· | ···················· | ···················· | ···················· | ···················· |
| Z | 4752 | hCoV-19/England/QEUH-B7E011/2020\|EPI_ISL_741934\|20 | ···················· | ···················· | ···················· | ···················· | ···················· | ···················· |
| Z | 4753 | hCoV-19/England/QEUH-B7E07B/2020\|EPI_ISL_741935\|20 | ···················· | ···················· | ···················· | ···················· | ···················· | ···················· |
| Z | 4754 | hCoV-19/England/SHEF-10C56F3/2020\|EPI_ISL_741936\|2 | ···················· | ···················· | ···················· | ···················· | ···················· | ···················· |
| Z | 4755 | hCoV-19/England/SHEF-10C5875/2020\|EPI_ISL_741937\|2 | ···················· | ···················· | ···················· | ···················· | ···················· | ···················· |
| Z | 4756 | hCoV-19/England/SHEF-10C7451/2020\|EPI_ISL_741947\|2 | ···················· | ···················· | ···················· | ···················· | ···················· | ···················· |
| Z | 4757 | hCoV-19/England/SHEF-10C7794/2020\|EPI_ISL_741948\|2 | ···················· | ···················· | ···················· | ···················· | ···················· | ···················· |
| Z | 4758 | hCoV-19/England/SHEF-10C79CB/2020\|EPI_ISL_741949\|2 | ···················· | ···················· | ···················· | ···················· | ·····T·············· | ···················· |
| Z | 4759 | hCoV-19/England/SHEF-10C803E/2020\|EPI_ISL_741950\|2 | ···················· | ···················· | ···················· | ···················· | ·····T·············· | ···················· |
| Z | 4760 | hCoV-19/England/SHEF-10C8872/2020\|EPI_ISL_741954\|2 | ···················· | ···················· | ···················· | ···················· | ···················· | ··A················· |
| Z | 4761 | hCoV-19/England/SHEF-10C89D9/2020\|EPI_ISL_741956\|2 | ···················· | ···················· | ···················· | ···················· | ···················· | ···················· |
| Z | 4762 | hCoV-19/England/SHEF-10C9422/2020\|EPI_ISL_741964\|2 | ···················· | ···················· | ···················· | ···················· | ···················· | ···················· |
| Z | 4763 | hCoV-19/England/SHEF-10C9F12/2020\|EPI_ISL_741967\|2 | ···················· | ···················· | ···················· | ···················· | ···················· | ···················· |
| Z | 4764 | hCoV-19/England/SHEF-10CA4A9/2020\|EPI_ISL_741969\|2 | ···················· | ···················· | ···················· | ···················· | ···················· | ···················· |
| Z | 4765 | hCoV-19/England/SHEF-10CAE7E/2020\|EPI_ISL_741976\|2 | ···················· | ···················· | ···················· | ···················· | ···················· | ···················· |
| Z | 4766 | hCoV-19/England/SHEF-10CB341/2020\|EPI_ISL_741978\|2 | ···················· | ···················· | ···················· | ···················· | ···················· | ···················· |
| Z | 4767 | hCoV-19/England/SHEF-10CBA2E/2020\|EPI_ISL_741981\|2 | ···················· | ···················· | ···················· | ···················· | ···················· | ···················· |
| Z | 4768 | hCoV-19/England/SHEF-10CBFD4/2020\|EPI_ISL_741986\|2 | ···················· | ···················· | ···················· | ···················· | ···················· | ···················· |
| Z | 4769 | hCoV-19/England/SHEF-10CC87E/2020\|EPI_ISL_741994\|2 | ···················· | ···················· | ···················· | ···················· | ···················· | ···················· |
| Z | 4770 | hCoV-19/England/SHEF-10CCC72/2020\|EPI_ISL_741995\|2 | ···················· | ···················· | ···················· | ···················· | ···················· | ···················· |
| Z | 4771 | hCoV-19/Wales/PHWC-4A3539/2020\|EPI_ISL_744108\|2020 | ···················· | ···················· | ···················· | ···················· | ···················· | ···················· |
| Z | 4772 | hCoV-19/Wales/PHWC-4A3548/2020\|EPI_ISL_744109\|2020 | ···················· | ···················· | ···················· | ···················· | ···················· | ···················· |
| Z | 4773 | hCoV-19/Wales/PHWC-4A3557/2020\|EPI_ISL_744110\|2020 | ···················· | ···················· | ···················· | ···················· | ···················· | ···················· |
| Z | 4774 | hCoV-19/Wales/PHWC-4A3566/2020\|EPI_ISL_744111\|2020 | ···················· | ···················· | ···················· | ···················· | ···················· | ···················· |

| | | | | | | | | |
|---|---|---|---|---|---|---|---|---|
| Z | 4775 | hCoV-19/Wales/PHWC-4A3575/2020\|EPI_ISL_744112\|2020 | ···················· | ···················· | ···················· | ···················· | ···················· | ···················· |
| Z | 4776 | hCoV-19/Wales/PHWC-4A3584/2020\|EPI_ISL_744113\|2020 | ···················· | ···················· | ···················· | ···················· | ···················· | ···················· |
| Z | 4777 | hCoV-19/Wales/PHWC-4A3593/2020\|EPI_ISL_744114\|2020 | ···················· | ···················· | ···················· | ···················· | ···················· | ···················· |
| Z | 4778 | hCoV-19/Wales/PHWC-4A35C0/2020\|EPI_ISL_744115\|2020 | ···················· | ···················· | ···················· | ···················· | ···················· | ···················· |
| Z | 4779 | hCoV-19/Wales/PHWC-4A35FD/2020\|EPI_ISL_744117\|2020 | ···················· | ···················· | ···················· | ···················· | ···················· | ···················· |
| Z | 4780 | hCoV-19/Wales/PHWC-4A3609/2020\|EPI_ISL_744118\|2020 | ···················· | ···················· | ···················· | ···················· | ···················· | ···················· |
| Z | 4781 | hCoV-19/Wales/PHWC-4A3618/2020\|EPI_ISL_744119\|2020 | ···················· | ···················· | ···················· | ···················· | ···················· | ···················· |
| Z | 4782 | hCoV-19/Wales/PHWC-4A3663/2020\|EPI_ISL_744121\|2020 | ···················· | ···················· | ···················· | ···················· | ···················· | ···················· |
| Z | 4783 | hCoV-19/Wales/PHWC-4A36DC/2020\|EPI_ISL_744126\|2020 | ···················· | ···················· | ···················· | ···················· | ···················· | ···················· |
| Z | 4784 | hCoV-19/Wales/PHWC-4A36EB/2020\|EPI_ISL_744127\|2020 | ···················· | ···················· | ···················· | ···················· | ···················· | ···················· |
| Z | 4785 | hCoV-19/Wales/PHWC-4A3706/2020\|EPI_ISL_744129\|2020 | ···················· | ···················· | ···················· | ···················· | ···················· | ···················· |
| Z | 4786 | hCoV-19/Luxembourg/LNS4860915/2020\|EPI_ISL_744150\| | ···················· | ···················· | ···················· | ···················· | ···················· | ···················· |
| Z | 4787 | hCoV-19/Luxembourg/LNS4874157/2020\|EPI_ISL_744152\| | ···················· | ···················· | ···················· | ···················· | ···················· | ···················· |
| Z | 4788 | hCoV-19/Luxembourg/LNS4889674/2020\|EPI_ISL_744153\| | ···················· | ···················· | ···················· | ···················· | ···················· | ···················· |
| Z | 4789 | hCoV-19/Luxembourg/LNS4891196/2020\|EPI_ISL_744154\| | ···················· | ···················· | ···················· | ···················· | ···················· | ···················· |
| Z | 4790 | hCoV-19/Luxembourg/LNS4905512/2020\|EPI_ISL_744155\| | ···················· | ···················· | ···················· | ···················· | ···················· | ···················· |
| Z | 4791 | hCoV-19/Luxembourg/LNS4906867/2020\|EPI_ISL_744156\| | ···················· | ···················· | ···················· | ···················· | ···················· | ···················· |
| Z | 4792 | hCoV-19/Luxembourg/LNS4910384/2020\|EPI_ISL_744157\| | ···················· | ···················· | ···················· | ···················· | ···················· | ···················· |
| Z | 4793 | hCoV-19/Luxembourg/LNS4916526/2020\|EPI_ISL_744158\| | ···················· | ···················· | ···················· | ···················· | ···················· | ···················· |
| Z | 4794 | hCoV-19/Luxembourg/LNS4916842/2020\|EPI_ISL_744159\| | ···················· | ···················· | ···················· | ···················· | ···················· | ···················· |
| Z | 4795 | hCoV-19/Luxembourg/LNS4918228/2020\|EPI_ISL_744160\| | ···················· | ···················· | ···················· | ···················· | ···················· | ···················· |
| Z | 4796 | hCoV-19/Luxembourg/LNS4922755/2020\|EPI_ISL_744161\| | ···················· | ···················· | ···················· | ···················· | ···················· | ···················· |
| Z | 4797 | hCoV-19/Luxembourg/LNS4925177/2020\|EPI_ISL_744162\| | ···················· | ···················· | ···················· | ···················· | ···················· | ···················· |
| Z | 4798 | hCoV-19/Luxembourg/LNS4933943/2020\|EPI_ISL_744163\| | ···················· | ···················· | ···················· | ···················· | ···················· | ···················· |
| Z | 4799 | hCoV-19/Luxembourg/LNS4957650/2020\|EPI_ISL_744164\| | ···················· | ···················· | ···················· | ···················· | ···················· | ···················· |
| Z | 4800 | hCoV-19/Luxembourg/LNS4969344/2020\|EPI_ISL_744165\| | ···················· | ···················· | ···················· | ···················· | ···················· | ···················· |
| Z | 4801 | hCoV-19/Luxembourg/LNS4978035/2020\|EPI_ISL_744166\| | ···················· | ·A·················· | ···················· | ···················· | ···················· | ···················· |
| Z | 4802 | hCoV-19/Luxembourg/LNS4978347/2020\|EPI_ISL_744167\| | ···················· | ···················· | ···················· | ···················· | ···················· | ···················· |
| Z | 4803 | hCoV-19/Luxembourg/LNS4979532/2020\|EPI_ISL_744168\| | ···················· | ···················· | ···················· | ···················· | ···················· | ···················· |
| Z | 4804 | hCoV-19/Luxembourg/LNS4988837/2020\|EPI_ISL_744169\| | ···················· | ···················· | ···················· | ···················· | ···················· | ···················· |
| Z | 4805 | hCoV-19/Luxembourg/LNS4989532/2020\|EPI_ISL_744170\| | ···················· | ···················· | ···················· | ···················· | ···················· | ···················· |
| Z | 4806 | hCoV-19/Luxembourg/LNS4993901/2020\|EPI_ISL_744171\| | ···················· | ···················· | ···················· | ···················· | ···················· | ···················· |
| Z | 4807 | hCoV-19/Luxembourg/LNS4996262/2020\|EPI_ISL_744172\| | ···················· | ···················· | ···················· | ···················· | ···················· | ···················· |
| Z | 4808 | hCoV-19/Luxembourg/LNS5011261/2020\|EPI_ISL_744174\| | ···················· | ···················· | ···················· | ···················· | ······T············· | ···················· |
| Z | 4809 | hCoV-19/Luxembourg/LNS5015530/2020\|EPI_ISL_744175\| | ···················· | ···················· | ···················· | ···················· | ···················· | ···················· |
| Z | 4810 | hCoV-19/Luxembourg/LNS5027174/2020\|EPI_ISL_744176\| | ···················· | ···················· | ···················· | ···················· | ···················· | ···················· |
| Z | 4811 | hCoV-19/Luxembourg/LNS5051188/2020\|EPI_ISL_744177\| | ···················· | ···················· | ···················· | ···················· | ···················· | ···················· |
| Z | 4812 | hCoV-19/Luxembourg/LNS5051676/2020\|EPI_ISL_744178\| | ···················· | ···················· | ···················· | ···················· | ···················· | ···················· |
| Z | 4813 | hCoV-19/Luxembourg/LNS5053760/2020\|EPI_ISL_744179\| | ···················· | ···················· | ···················· | ···················· | ···················· | ···················· |
| Z | 4814 | hCoV-19/Luxembourg/LNS5064005/2020\|EPI_ISL_744180\| | ···················· | ···················· | ···················· | ···················· | ···················· | ···················· |
| Z | 4815 | hCoV-19/Luxembourg/LNS5070809/2020\|EPI_ISL_744181\| | ···················· | ···················· | ···················· | ···················· | ···················· | ···················· |
| Z | 4816 | hCoV-19/Luxembourg/LNS5082645/2020\|EPI_ISL_744182\| | ···················· | ···················· | ···················· | ···················· | ···················· | ···················· |
| Z | 4817 | hCoV-19/Luxembourg/LNS5091746/2020\|EPI_ISL_744183\| | ···················· | ···················· | ···················· | ···················· | ···················· | ···················· |
| Z | 4818 | hCoV-19/Luxembourg/LNS5100136/2020\|EPI_ISL_744184\| | ···················· | ···················· | ···················· | ···················· | ···················· | ···················· |
| Z | 4819 | hCoV-19/Luxembourg/LNS5109849/2020\|EPI_ISL_744185\| | ···················· | ···················· | ···················· | ···················· | ···················· | ···················· |
| Z | 4820 | hCoV-19/Luxembourg/LNS5111129/2020\|EPI_ISL_744186\| | ···················· | ···················· | ···················· | ···················· | ···················· | ···················· |
| Z | 4821 | hCoV-19/Luxembourg/LNS5121437/2020\|EPI_ISL_744187\| | ···················· | ···················· | ···················· | ···················· | ···················· | ···················· |
| Z | 4822 | hCoV-19/Luxembourg/LNS5124167/2020\|EPI_ISL_744188\| | ···················· | ···················· | ···················· | ···················· | ···················· | ···················· |
| Z | 4823 | hCoV-19/Luxembourg/LNS5141929/2020\|EPI_ISL_744190\| | ···················· | ···················· | ···················· | ···················· | ···················· | ···················· |
| Z | 4824 | hCoV-19/Luxembourg/LNS5146889/2020\|EPI_ISL_744191\| | ···················· | ···················· | ···················· | ···················· | ···················· | ···················· |
| Z | 4825 | hCoV-19/Luxembourg/LNS5151925/2020\|EPI_ISL_744192\| | ···················· | ···················· | ···················· | ···················· | ···················· | ···················· |
| Z | 4826 | hCoV-19/Luxembourg/LNS5156934/2020\|EPI_ISL_744193\| | ···················· | ···················· | ···················· | ···················· | ···················· | ···················· |
| Z | 4827 | hCoV-19/Luxembourg/LNS5157058/2020\|EPI_ISL_744194\| | ···················· | ···················· | ···················· | ···················· | ···················· | ···················· |
| Z | 4828 | hCoV-19/Luxembourg/LNS5163154/2020\|EPI_ISL_744195\| | ···················· | ···················· | ···················· | ···················· | ···················· | ···················· |
| Z | 4829 | hCoV-19/Luxembourg/LNS5175625/2020\|EPI_ISL_744196\| | ···················· | ···················· | ···················· | ···················· | ···················· | ···················· |
| Z | 4830 | hCoV-19/Luxembourg/LNS5178768/2020\|EPI_ISL_744197\| | ···················· | ···················· | ···················· | ···················· | ···················· | ···················· |
| Z | 4831 | hCoV-19/Luxembourg/LNS5185009/2020\|EPI_ISL_744198\| | ···················· | ···················· | ···················· | ···················· | ···················· | ···················· |
| Z | 4832 | hCoV-19/Luxembourg/LNS5186473/2020\|EPI_ISL_744199\| | ···················· | ···················· | ···················· | ···················· | ···················· | ···················· |
| Z | 4833 | hCoV-19/USA/NY-NYCPHL-001524/2020\|EPI_ISL_745439\|2 | ···················· | ···················· | ···················· | ···················· | ···················· | ···················· |
| Z | 4834 | hCoV-19/USA/NY-NYCPHL-001525/2020\|EPI_ISL_745440\|2 | ···················· | ···················· | ···················· | ···················· | ···················· | ···················· |
| Z | 4835 | hCoV-19/USA/NY-NYCPHL-001205/2020\|EPI_ISL_745441\|2 | ···················· | ·A·················· | ···················· | ···················· | ···················· | ···················· |
| Z | 4836 | hCoV-19/USA/NY-NYCPHL-001206/2020\|EPI_ISL_745442\|2 | ···················· | ···················· | ···················· | ···················· | ···················· | ···················· |
| Z | 4837 | hCoV-19/USA/NY-NYCPHL-001207/2020\|EPI_ISL_745443\|2 | ···················· | ···················· | ···················· | ···················· | ···················· | ···················· |
| Z | 4838 | hCoV-19/USA/NY-NYCPHL-001209/2020\|EPI_ISL_745444\|2 | ···················· | ···················· | ···················· | ···················· | ···················· | ···················· |
| Z | 4839 | hCoV-19/USA/NY-NYCPHL-001526/2020\|EPI_ISL_745445\|2 | ···················· | ······A············· | ···················· | ···················· | ···················· | ···················· |
| Z | 4840 | hCoV-19/USA/NY-NYCPHL-001527/2020\|EPI_ISL_745446\|2 | ···················· | ···················· | ···················· | ···················· | ···················· | ···················· |
| Z | 4841 | hCoV-19/USA/NY-NYCPHL-001528/2020\|EPI_ISL_745447\|2 | ···················· | ···················· | ···················· | ···················· | ···················· | ···················· |
| Z | 4842 | hCoV-19/USA/NY-NYCPHL-001529/2020\|EPI_ISL_745448\|2 | ···················· | ···················· | ···················· | ···················· | ···················· | ···················· |
| Z | 4843 | hCoV-19/USA/NY-NYCPHL-001530/2020\|EPI_ISL_745449\|2 | ···················· | ·A·················· | ···················· | ···················· | ···················· | ···················· |
| Z | 4844 | hCoV-19/USA/NY-NYCPHL-001532/2020\|EPI_ISL_745450\|2 | ···················· | ···················· | ···················· | ···················· | ···················· | ···················· |
| Z | 4845 | hCoV-19/USA/NY-NYCPHL-001533/2020\|EPI_ISL_745451\|2 | ···················· | ···················· | ···················· | ···················· | ···················· | ···················· |
| Z | 4846 | hCoV-19/USA/NY-NYCPHL-001534/2020\|EPI_ISL_745452\|2 | ···················· | ·A·················· | ···················· | ···················· | ···················· | ···················· |
| Z | 4847 | hCoV-19/USA/NY-NYCPHL-001541/2020\|EPI_ISL_745457\|2 | ···················· | ···················· | ···················· | ···················· | ···················· | ···················· |
| Z | 4848 | hCoV-19/USA/NY-NYCPHL-001542/2020\|EPI_ISL_745458\|2 | ···················· | ···················· | ···················· | ···················· | ···················· | ···················· |
| Z | 4849 | hCoV-19/Chile/RM-208464/2020\|EPI_ISL_746770\|2020-1 | ···················· | ···················· | ···················· | ···················· | ···················· | ···················· |
| Z | 4850 | hCoV-19/Chile/RM-208466/2020\|EPI_ISL_746771\|2020-1 | ···················· | ···················· | ···················· | ···················· | ···················· | ···················· |
| Z | 4851 | hCoV-19/Chile/RM-208555/2020\|EPI_ISL_746772\|2020-1 | ···················· | ···················· | ···················· | ···················· | ···················· | ···················· |
| Z | 4852 | hCoV-19/Chile/RM-208669/2020\|EPI_ISL_746773\|2020-1 | ···················· | ···················· | ···················· | ···················· | ···················· | ···················· |
| Z | 4853 | hCoV-19/Chile/RM-72426/2020\|EPI_ISL_746774\|2020-05 | ···················· | ···················· | ···················· | ···················· | ···················· | ···················· |
| Z | 4854 | hCoV-19/Chile/RM-72428/2020\|EPI_ISL_746775\|2020-05 | ···················· | ···················· | ···················· | ···················· | ···················· | ···················· |

| | | | | | | | | |
|---|---|---|---|---|---|---|---|---|
| Z | 4855 | hCoV-19/Chile/RM-72433/2020\|EPI_ISL_746776\|2020-05 | ···················· | ···················· | ····················· | ···················· | ···················· | ···················· |
| Z | 4856 | hCoV-19/Chile/RM-72434/2020\|EPI_ISL_746777\|2020-05 | ···················· | ···················· | ····················· | ···················· | ···················· | ···················· |
| Z | 4857 | hCoV-19/Chile/RM-72815/2020\|EPI_ISL_746778\|2020-05 | ···················· | ···················· | ····················· | ···················· | ···················· | ···················· |
| Z | 4858 | hCoV-19/Chile/RM-72817/2020\|EPI_ISL_746779\|2020-05 | ···················· | ···················· | ····················· | ···················· | ···················· | ···················· |
| Z | 4859 | hCoV-19/Chile/RM-72820/2020\|EPI_ISL_746780\|2020-05 | ···················· | ···················· | ····················· | ···················· | ···················· | ···················· |
| Z | 4860 | hCoV-19/Chile/RM-72821/2020\|EPI_ISL_746781\|2020-05 | ···················· | ···················· | ····················· | ···················· | ···················· | ···················· |
| Z | 4861 | hCoV-19/Chile/RM-72843/2020\|EPI_ISL_746782\|2020-05 | ···················· | ···················· | ····················· | ···················· | ···················· | ···················· |
| Z | 4862 | hCoV-19/Chile/RM-72846/2020\|EPI_ISL_746783\|2020-05 | ···················· | ···················· | ····················· | ···················· | ···················· | ···················· |
| Z | 4863 | hCoV-19/Chile/RM-73246/2020\|EPI_ISL_746788\|2020-05 | ···················· | ···················· | ····················· | ···················· | ···················· | ···················· |
| Z | 4864 | hCoV-19/Chile/RM-73253/2020\|EPI_ISL_746789\|2020-05 | ···················· | ···················· | ····················· | ···················· | ···················· | ···················· |
| Z | 4865 | hCoV-19/USA/CA-CZB-14132/2020\|EPI_ISL_739640\|2020- | ···················· | ···················· | ····················· | ···················· | ···················· | ···················· |
| Z | 4866 | hCoV-19/Canada/BC-BCCDC-7234/2020\|EPI_ISL_740626\|2 | ···················· | ···················· | ····················· | ···················· | ···················· | ···················· |
| Z | 4867 | hCoV-19/Canada/BC-BCCDC-7235/2020\|EPI_ISL_740627\|2 | ···················· | ···················· | ····················· | ···················· | ···················· | ···················· |
| Z | 4868 | hCoV-19/England/CAMC-B7A33A/2020\|EPI_ISL_741100\|20 | ···················· | ···················· | ····················· | ···················· | ···················· | ···················· |
| Z | 4869 | hCoV-19/England/CAMC-B7A349/2020\|EPI_ISL_741101\|20 | ···················· | ···················· | ····················· | ···················· | ···················· | ···················· |
| Z | 4870 | hCoV-19/England/CAMC-B7A358/2020\|EPI_ISL_741102\|20 | ···················· | ···················· | ····················· | ···················· | ···················· | ···················· |
| Z | 4871 | hCoV-19/England/CAMC-B7A376/2020\|EPI_ISL_741103\|20 | ···················· | ···················· | ····················· | ···················· | ···················· | ···················· |
| Z | 4872 | hCoV-19/England/CAMC-B7A3A3/2020\|EPI_ISL_741105\|20 | ···················· | ···················· | ····················· | ···················· | ···················· | ···················· |
| Z | 4873 | hCoV-19/England/CAMC-B7A3B2/2020\|EPI_ISL_741106\|20 | ···················· | ···················· | ····················· | ···················· | ···················· | ···················· |
| Z | 4874 | hCoV-19/England/CAMC-B7A3D0/2020\|EPI_ISL_741107\|20 | ···················· | ···················· | ····················· | ···················· | ···················· | ···················· |
| Z | 4875 | hCoV-19/England/CAMC-B7A40A/2020\|EPI_ISL_741108\|20 | ···················· | ···················· | ····················· | ···················· | ···················· | ···················· |
| Z | 4876 | hCoV-19/England/CAMC-B7A437/2020\|EPI_ISL_741110\|20 | ···················· | ·A·················· | ····················· | ···················· | ···················· | ··A················· |
| Z | 4877 | hCoV-19/England/CAMC-B7A455/2020\|EPI_ISL_741111\|20 | ···················· | ···················· | ····················· | ···················· | ···················· | ···················· |
| Z | 4878 | hCoV-19/England/CAMC-B7A464/2020\|EPI_ISL_741112\|20 | ···················· | ···················· | ····················· | ···················· | ···················· | ···················· |
| Z | 4879 | hCoV-19/England/CAMC-B7A473/2020\|EPI_ISL_741113\|20 | ···················· | ···················· | ····················· | ···················· | ···················· | ···················· |
| Z | 4880 | hCoV-19/England/CAMC-B7A491/2020\|EPI_ISL_741114\|20 | ···················· | ···················· | ····················· | ···················· | ···················· | ···················· |
| Z | 4881 | hCoV-19/England/CAMC-B7A4DD/2020\|EPI_ISL_741117\|20 | ···················· | ···················· | ····················· | ···················· | ···················· | ···················· |
| Z | 4882 | hCoV-19/England/CAMC-B7A4EC/2020\|EPI_ISL_741118\|20 | ···················· | ···················· | ····················· | ···················· | ···················· | ···················· |
| Z | 4883 | hCoV-19/England/CAMC-B7A4FB/2020\|EPI_ISL_741119\|20 | ···················· | ···················· | ····················· | ···················· | ···················· | ···················· |
| Z | 4884 | hCoV-19/England/CAMC-B7A516/2020\|EPI_ISL_741120\|20 | ···················· | ···················· | ····················· | ···················· | ······T············· | ···················· |
| Z | 4885 | hCoV-19/England/CAMC-B7A525/2020\|EPI_ISL_741121\|20 | ···················· | ···················· | ····················· | ···················· | ···················· | ···················· |
| Z | 4886 | hCoV-19/England/CAMC-B7A552/2020\|EPI_ISL_741122\|20 | ···················· | ···················· | ····················· | ···················· | ···················· | ···················· |
| Z | 4887 | hCoV-19/England/CAMC-B7A561/2020\|EPI_ISL_741123\|20 | ···················· | ···················· | ····················· | ···················· | ···················· | ···················· |
| Z | 4888 | hCoV-19/England/CAMC-B7A570/2020\|EPI_ISL_741124\|20 | ···················· | ···················· | ····················· | ···················· | ···················· | ···················· |
| Z | 4889 | hCoV-19/England/CAMC-B7A58F/2020\|EPI_ISL_741125\|20 | ···················· | ···················· | ····················· | ···················· | ···················· | ···················· |
| Z | 4890 | hCoV-19/England/CAMC-B7A5CB/2020\|EPI_ISL_741127\|20 | ···················· | ···················· | ····················· | ···················· | ···················· | ···················· |
| Z | 4891 | hCoV-19/England/CAMC-B7A5DA/2020\|EPI_ISL_741128\|20 | ···················· | ···················· | ····················· | ···················· | ···················· | ···················· |
| Z | 4892 | hCoV-19/England/CAMC-B7A65F/2020\|EPI_ISL_741131\|20 | ···················· | ···················· | ····················· | ···················· | ······T············· | ···················· |
| Z | 4893 | hCoV-19/England/CAMC-B7A66E/2020\|EPI_ISL_741132\|20 | ···················· | ···················· | ····················· | ···················· | ···················· | ···················· |
| Z | 4894 | hCoV-19/England/CAMC-B7A6B9/2020\|EPI_ISL_741133\|20 | ········T··········· | ···················· | ····················· | ···················· | ···················· | ···················· |
| Z | 4895 | hCoV-19/England/CAMC-B7A6D7/2020\|EPI_ISL_741134\|20 | ···················· | ···················· | ····················· | ···················· | ······T············· | ···················· |
| Z | 4896 | hCoV-19/England/CAMC-B7A710/2020\|EPI_ISL_741136\|20 | ···················· | ···················· | ····················· | ···················· | ···················· | ···················· |
| Z | 4897 | hCoV-19/England/CAMC-B7A74D/2020\|EPI_ISL_741137\|20 | ···················· | ·A·················· | ····················· | ···················· | ···················· | ···················· |
| Z | 4898 | hCoV-19/England/CAMC-B7A789/2020\|EPI_ISL_741138\|20 | ···················· | ···················· | ····················· | ···················· | ···················· | ···················· |
| Z | 4899 | hCoV-19/England/CAMC-B7A798/2020\|EPI_ISL_741139\|20 | ···················· | ···················· | ····················· | ···················· | ···················· | ···················· |
| Z | 4900 | hCoV-19/England/CAMC-B7A7A7/2020\|EPI_ISL_741140\|20 | ···················· | ···················· | ····················· | ···················· | ···················· | ···················· |
| Z | 4901 | hCoV-19/England/CAMC-B7A7B6/2020\|EPI_ISL_741141\|20 | ···················· | ···················· | ····················· | ···················· | ···················· | ···················· |
| Z | 4902 | hCoV-19/England/CAMC-B7A7D4/2020\|EPI_ISL_741142\|20 | ···················· | ············C······· | ····················· | ···················· | ···················· | ···················· |
| Z | 4903 | hCoV-19/England/CAMC-B7A7E3/2020\|EPI_ISL_741143\|20 | ···················· | ···················· | ····················· | ···················· | ···················· | ···················· |
| Z | 4904 | hCoV-19/England/CAMC-B7A7F2/2020\|EPI_ISL_741144\|20 | ···················· | ···················· | ····················· | ···················· | ···················· | ···················· |
| Z | 4905 | hCoV-19/England/CAMC-B7A80E/2020\|EPI_ISL_741145\|20 | ···················· | ···················· | ····················· | ···················· | ···················· | ···················· |
| Z | 4906 | hCoV-19/England/CAMC-B7A83B/2020\|EPI_ISL_741147\|20 | ···················· | ···················· | ····················· | ···················· | ···················· | ···················· |
| Z | 4907 | hCoV-19/England/CAMC-B7A84A/2020\|EPI_ISL_741148\|20 | ········T··········· | ···················· | ····················· | ···················· | ···················· | ···················· |
| Z | 4908 | hCoV-19/England/CAMC-B7A868/2020\|EPI_ISL_741149\|20 | ···················· | ···················· | ····················· | ···················· | ···················· | ···················· |
| Z | 4909 | hCoV-19/England/CAMC-B7A895/2020\|EPI_ISL_741150\|20 | ···················· | ···················· | ····················· | ···················· | ···················· | ···················· |
| Z | 4910 | hCoV-19/England/CAMC-B7A8D1/2020\|EPI_ISL_741151\|20 | ···················· | ···················· | ····················· | ···················· | ···················· | ···················· |
| Z | 4911 | hCoV-19/England/CAMC-B7A938/2020\|EPI_ISL_741152\|20 | ···················· | ···················· | ····················· | ···················· | ···················· | ···················· |
| Z | 4912 | hCoV-19/England/CAMC-B7A947/2020\|EPI_ISL_741153\|20 | ···················· | ···················· | ····················· | ···················· | ···················· | ···················· |
| Z | 4913 | hCoV-19/England/CAMC-B7AA26/2020\|EPI_ISL_741154\|20 | ···················· | ···················· | ····················· | ···················· | ···················· | ···················· |
| Z | 4914 | hCoV-19/England/CAMC-B7AA44/2020\|EPI_ISL_741155\|20 | ···················· | ···················· | ····················· | ···················· | ···················· | ···················· |
| Z | 4915 | hCoV-19/England/CAMC-B7AA9F/2020\|EPI_ISL_741156\|20 | ···················· | ···················· | ····················· | ···················· | ···················· | ··A················· |
| Z | 4916 | hCoV-19/England/CAMC-B7AABD/2020\|EPI_ISL_741158\|20 | ···················· | ···················· | ····················· | ···················· | ···················· | ···················· |
| Z | 4917 | hCoV-19/England/CAMC-B7AACC/2020\|EPI_ISL_741159\|20 | ···················· | ···················· | ····················· | ···················· | ···················· | ···················· |
| Z | 4918 | hCoV-19/England/CAMC-B7AB23/2020\|EPI_ISL_741161\|20 | ···················· | ···················· | ····················· | ···················· | ···················· | ···················· |
| Z | 4919 | hCoV-19/England/CAMC-B7AB32/2020\|EPI_ISL_741162\|20 | ···················· | ···················· | ····················· | ···················· | ···················· | ···················· |
| Z | 4920 | hCoV-19/England/CAMC-B7AB41/2020\|EPI_ISL_741163\|20 | ···················· | ···················· | ····················· | ···················· | ···················· | ···················· |
| Z | 4921 | hCoV-19/England/CAMC-B7AB50/2020\|EPI_ISL_741164\|20 | ···················· | ···················· | ····················· | ···················· | ···················· | ···················· |
| Z | 4922 | hCoV-19/England/CAMC-B7AB8D/2020\|EPI_ISL_741166\|20 | ···················· | ···················· | ····················· | ···················· | ···················· | ···················· |
| Z | 4923 | hCoV-19/England/CAMC-B7ABC9/2020\|EPI_ISL_741169\|20 | ···················· | ···················· | ····················· | ···················· | ···················· | ···················· |
| Z | 4924 | hCoV-19/England/CAMC-B7ABD8/2020\|EPI_ISL_741170\|20 | ···················· | ···················· | ····················· | ···················· | ···················· | ···················· |
| Z | 4925 | hCoV-19/England/CAMC-B7AD1E/2020\|EPI_ISL_741171\|20 | ···················· | ···················· | ····················· | ···················· | ···················· | ···················· |
| Z | 4926 | hCoV-19/England/CAMC-B7AD4B/2020\|EPI_ISL_741173\|20 | ···················· | ···················· | ····················· | ···················· | ···················· | ···················· |
| Z | 4927 | hCoV-19/England/CAMC-B7AD87/2020\|EPI_ISL_741175\|20 | ···················· | ···················· | ····················· | ···················· | ······T············· | ···················· |
| Z | 4928 | hCoV-19/England/CAMC-B7B515/2020\|EPI_ISL_741176\|20 | ···················· | ···················· | ····················· | ···················· | ···················· | ···················· |
| Z | 4929 | hCoV-19/England/CAMC-B7B524/2020\|EPI_ISL_741177\|20 | ···················· | ···················· | ····················· | ···················· | ···················· | ···················· |
| Z | 4930 | hCoV-19/England/CAMC-B7B533/2020\|EPI_ISL_741178\|20 | ···················· | ·A·················· | ····················· | ···················· | ···················· | ···················· |
| Z | 4931 | hCoV-19/England/CAMC-B7B560/2020\|EPI_ISL_741179\|20 | ···················· | ·A·················· | ····················· | ···················· | ···················· | ··A················· |
| Z | 4932 | hCoV-19/England/CAMC-B7B57F/2020\|EPI_ISL_741180\|20 | ···················· | ···················· | ····················· | ···················· | ···················· | ···················· |
| Z | 4933 | hCoV-19/England/CAMC-B7B58E/2020\|EPI_ISL_741181\|20 | ···················· | ···················· | ····················· | ···················· | ···················· | ···················· |
| Z | 4934 | hCoV-19/England/CAMC-B7B5BB/2020\|EPI_ISL_741183\|20 | ···················· | ·A·················· | ····················· | ···················· | ···················· | ···················· |

| | | | | | | | | |
|---|---|---|---|---|---|---|---|---|
| Z | 4935 | hCoV-19/England/CAMC-B7B5CA/2020\|EPI_ISL_741184\|20 | ···················· | ···················· | ···················· | ···················· | ···················· | ···················· |
| Z | 4936 | hCoV-19/England/CAMC-B7B5E8/2020\|EPI_ISL_741185\|20 | ···················· | ···················· | ···················· | ···················· | ···················· | ···················· |
| Z | 4937 | hCoV-19/England/LOND-1266C4B/2020\|EPI_ISL_741189\|2 | ···················· | ·A·················· | ···················· | ···················· | ···················· | ···A················ |
| Z | 4938 | hCoV-19/England/LOND-1266C69/2020\|EPI_ISL_741191\|2 | ···················· | ···················· | ···················· | ···················· | ···················· | ···················· |
| Z | 4939 | hCoV-19/England/LOND-1266CD2/2020\|EPI_ISL_741196\|2 | ···················· | ···················· | ···················· | ···················· | ···················· | ···················· |
| Z | 4941 | hCoV-19/Wales/PHWC-49B8D1/2020\|EPI_ISL_742401\|2020 | ···················· | ···················· | ···················· | ···················· | ···················· | ···················· |
| Z | 4943 | hCoV-19/Wales/PHWC-49B90B/2020\|EPI_ISL_742403\|2020 | ···················· | ···················· | ···················· | ···················· | ···················· | ···················· |
| Z | 4945 | hCoV-19/Wales/PHWC-49B965/2020\|EPI_ISL_742407\|2020 | ···················· | ···················· | ···················· | ···················· | ···················· | ···················· |
| Z | 4946 | hCoV-19/Wales/PHWC-49B992/2020\|EPI_ISL_742409\|2020 | ···················· | ···················· | ···················· | ···················· | ···················· | ···················· |
| Z | 4950 | hCoV-19/Wales/PHWC-49B9DE/2020\|EPI_ISL_742413\|2020 | ···················· | ···················· | ···················· | ···················· | ···················· | ···················· |
| Z | 4953 | hCoV-19/Wales/PHWC-49BA26/2020\|EPI_ISL_742417\|2020 | ···················· | ···················· | ···················· | ···················· | ···················· | ···················· |
| Z | 4955 | hCoV-19/Wales/PHWC-49BA44/2020\|EPI_ISL_742419\|2020 | ···················· | ···················· | ···················· | ···················· | ···················· | ···················· |
| Z | 4958 | hCoV-19/Wales/PHWC-49BA80/2020\|EPI_ISL_742423\|2020 | ···················· | ···················· | ···················· | ···················· | ···················· | ···················· |
| Z | 4959 | hCoV-19/Wales/PHWC-49BA9F/2020\|EPI_ISL_742424\|2020 | ···················· | ···················· | ···················· | ···················· | ···················· | ···················· |
| Z | 4961 | hCoV-19/Wales/PHWC-49BACC/2020\|EPI_ISL_742427\|2020 | ···················· | ···················· | ···················· | ···················· | ···················· | ···················· |
| Z | 4962 | hCoV-19/Wales/PHWC-49BADB/2020\|EPI_ISL_742428\|2020 | ···················· | ···················· | ···················· | ···················· | ···················· | ···················· |
| Z | 4965 | hCoV-19/Wales/PHWC-49BB14/2020\|EPI_ISL_742432\|2020 | ···················· | ···················· | ···················· | ···················· | ···················· | ···················· |
| Z | 4966 | hCoV-19/Wales/PHWC-49BB23/2020\|EPI_ISL_742433\|2020 | ···················· | ···················· | ···················· | ···················· | ···················· | ···················· |
| Z | 4968 | hCoV-19/Wales/PHWC-49BB41/2020\|EPI_ISL_742435\|2020 | ···················· | ···················· | ···················· | ···················· | ···················· | ···················· |
| Z | 4969 | hCoV-19/Wales/PHWC-49BB50/2020\|EPI_ISL_742436\|2020 | ···················· | ···················· | ···················· | ···················· | ···················· | ···················· |
| Z | 4971 | hCoV-19/Wales/PHWC-49BB8D/2020\|EPI_ISL_742439\|2020 | ···················· | ···················· | ···················· | ···················· | ···················· | ···················· |
| Z | 4973 | hCoV-19/Wales/PHWC-49BBBA/2020\|EPI_ISL_742441\|2020 | ···················· | ···················· | ···················· | ···················· | ···················· | ···················· |
| Z | 4977 | hCoV-19/Wales/PHWC-49BC11/2020\|EPI_ISL_742446\|2020 | ···················· | ···················· | ···················· | ···················· | ···················· | ···················· |
| Z | 4978 | hCoV-19/Wales/PHWC-49BC20/2020\|EPI_ISL_742447\|2020 | ···················· | ···················· | ···················· | ···················· | ···················· | ···················· |
| Z | 4979 | hCoV-19/Wales/PHWC-49BC3F/2020\|EPI_ISL_742448\|2020 | ···················· | ···················· | ···················· | ···················· | ···················· | ···················· |
| Z | 4981 | hCoV-19/Wales/PHWC-49BC5D/2020\|EPI_ISL_742450\|2020 | ···················· | ···················· | ···················· | ···················· | ···················· | ···················· |
| Z | 4982 | hCoV-19/Wales/PHWC-49BC6C/2020\|EPI_ISL_742451\|2020 | ···················· | ···················· | ···················· | ···················· | ···················· | ···················· |
| Z | 4984 | hCoV-19/Wales/PHWC-49BC8A/2020\|EPI_ISL_742453\|2020 | ···················· | ···················· | ···················· | ···················· | ···················· | ···················· |
| Z | 4986 | hCoV-19/Wales/PHWC-49BCA8/2020\|EPI_ISL_742455\|2020 | ···················· | ···················· | ···················· | ···················· | ···················· | ···················· |
| Z | 4987 | hCoV-19/Wales/PHWC-49BCC6/2020\|EPI_ISL_742456\|2020 | ···················· | ···················· | ···················· | ···················· | ···················· | ···················· |
| Z | 4989 | hCoV-19/Wales/PHWC-49BD1E/2020\|EPI_ISL_742459\|2020 | ···················· | ···················· | ···················· | ···················· | ···················· | ···················· |
| Z | 4990 | hCoV-19/Wales/PHWC-49BD5A/2020\|EPI_ISL_742462\|2020 | ···················· | ···················· | ···················· | ···················· | ···················· | ···················· |
| Z | 4991 | hCoV-19/Wales/PHWC-49BD78/2020\|EPI_ISL_742464\|2020 | ···················· | ···················· | ···················· | ···················· | ···················· | ···················· |
| Z | 4994 | hCoV-19/Wales/PHWC-49BDD2/2020\|EPI_ISL_742467\|2020 | ···················· | ···················· | ···················· | ···················· | ···················· | ···················· |
| Z | 4995 | hCoV-19/Wales/PHWC-49BDE1/2020\|EPI_ISL_742468\|2020 | ···················· | ···················· | ···················· | ···················· | ···················· | ···················· |
| Z | 4998 | hCoV-19/Wales/PHWC-49BE1B/2020\|EPI_ISL_742471\|2020 | ···················· | ···················· | ···················· | ···················· | ···················· | ···················· |
| Z | 4999 | hCoV-19/Wales/PHWC-49BE48/2020\|EPI_ISL_742472\|2020 | ···················· | ···················· | ···················· | ···················· | ···················· | ···················· |
| Z | 5001 | hCoV-19/Wales/PHWC-49BE66/2020\|EPI_ISL_742474\|2020 | ···················· | ···················· | ···················· | ···················· | ···················· | ···················· |
| Z | 5003 | hCoV-19/Wales/PHWC-49BE93/2020\|EPI_ISL_742477\|2020 | ···················· | ···················· | ···················· | ···················· | ···················· | ···················· |
| Z | 5004 | hCoV-19/Wales/PHWC-49BEB1/2020\|EPI_ISL_742478\|2020 | ···················· | ···················· | ···················· | ···················· | ···················· | ···················· |
| Z | 5005 | hCoV-19/Wales/PHWC-49BEDF/2020\|EPI_ISL_742479\|2020 | ···················· | ···················· | ···················· | ···················· | ···················· | ···················· |
| Z | 5007 | hCoV-19/Wales/PHWC-49BEFD/2020\|EPI_ISL_742481\|2020 | ···················· | ···················· | ···················· | ···················· | ···················· | ···················· |
| Z | 5009 | hCoV-19/Wales/PHWC-49BF18/2020\|EPI_ISL_742483\|2020 | ···················· | ···················· | ···················· | ···················· | ···················· | ····N··············· |
| Z | 5010 | hCoV-19/Wales/PHWC-49BF27/2020\|EPI_ISL_742484\|2020 | ···················· | ···················· | ···················· | ···················· | ···················· | ···················· |
| Z | 5012 | hCoV-19/Wales/PHWC-49BF63/2020\|EPI_ISL_742486\|2020 | ···················· | ···················· | ···················· | ···················· | ···················· | ···················· |
| Z | 5014 | hCoV-19/Wales/PHWC-49BF90/2020\|EPI_ISL_742488\|2020 | ···················· | ···················· | ···················· | ···················· | ···················· | ···················· |
| Z | 5017 | hCoV-19/Wales/PHWC-49BFDC/2020\|EPI_ISL_742492\|2020 | ···················· | ···················· | ···················· | ···················· | ···················· | ···················· |
| Z | 5018 | hCoV-19/Wales/PHWC-49BFEB/2020\|EPI_ISL_742493\|2020 | ···················· | ···················· | ···················· | ···················· | ···················· | ···················· |
| Z | 5020 | hCoV-19/Wales/PHWC-49C005/2020\|EPI_ISL_742495\|2020 | ···················· | ···················· | ···················· | ···················· | ···················· | ···················· |
| Z | 5021 | hCoV-19/Wales/PHWC-49C023/2020\|EPI_ISL_742496\|2020 | ···················· | ···················· | ···················· | ···················· | ···················· | ···················· |
| Z | 5022 | hCoV-19/Wales/PHWC-49C032/2020\|EPI_ISL_742497\|2020 | ···················· | ···················· | ···················· | ···················· | ···················· | ···················· |
| Z | 5025 | hCoV-19/Wales/PHWC-4A160B/2020\|EPI_ISL_743700\|2020 | ···················· | ···················· | ···················· | ···················· | ···················· | ···················· |
| Z | 5026 | hCoV-19/Wales/PHWC-4A161A/2020\|EPI_ISL_743701\|2020 | ···················· | ···················· | ···················· | ···················· | ···················· | ···················· |
| Z | 5028 | hCoV-19/Wales/PHWC-4A1638/2020\|EPI_ISL_743703\|2020 | ···················· | ···················· | ···················· | ···················· | ···················· | ···················· |
| Z | 5029 | hCoV-19/Wales/PHWC-4A1647/2020\|EPI_ISL_743704\|2020 | ···················· | ···················· | ···················· | ···················· | ···················· | ···················· |
| Z | 5030 | hCoV-19/Wales/PHWC-4A1656/2020\|EPI_ISL_743705\|2020 | ···················· | ···················· | ···················· | ···················· | ···················· | ···················· |
| Z | 5031 | hCoV-19/Wales/PHWC-4A1665/2020\|EPI_ISL_743706\|2020 | ···················· | ···················· | ···················· | ···················· | ···················· | ···················· |
| Z | 5032 | hCoV-19/Wales/PHWC-4A1674/2020\|EPI_ISL_743707\|2020 | ···················· | ···················· | ···················· | ···················· | ···················· | ···················· |
| Z | 5033 | hCoV-19/Wales/PHWC-4A1683/2020\|EPI_ISL_743708\|2020 | ···················· | ···················· | ···················· | ···················· | ···················· | ···················· |
| Z | 5035 | hCoV-19/Wales/PHWC-4A16DE/2020\|EPI_ISL_743712\|2020 | ···················· | ···················· | ···················· | ···················· | ···················· | ···················· |
| Z | 5036 | hCoV-19/Wales/PHWC-4A16ED/2020\|EPI_ISL_743713\|2020 | ···················· | ···················· | ···················· | ···················· | ···················· | ···················· |
| Z | 5037 | hCoV-19/Wales/PHWC-4A1717/2020\|EPI_ISL_743715\|2020 | ···················· | ···················· | ···················· | ···················· | ···················· | ···················· |
| Z | 5038 | hCoV-19/Wales/PHWC-4A1726/2020\|EPI_ISL_743716\|2020 | ···················· | ···················· | ···················· | ···················· | ···················· | ···················· |
| Z | 5039 | hCoV-19/Wales/PHWC-4A1735/2020\|EPI_ISL_743717\|2020 | ···················· | ···················· | ···················· | ···················· | ···················· | ···················· |
| Z | 5040 | hCoV-19/Wales/PHWC-4A1744/2020\|EPI_ISL_743718\|2020 | ···················· | ···················· | ···················· | ···················· | ···················· | ···················· |
| Z | 5041 | hCoV-19/Wales/PHWC-4A1762/2020\|EPI_ISL_743720\|2020 | ···················· | ···················· | ···················· | ···················· | ···················· | ···················· |
| Z | 5042 | hCoV-19/Wales/PHWC-4A1771/2020\|EPI_ISL_743721\|2020 | ···················· | ···················· | ···················· | ···················· | ···················· | ···················· |
| Z | 5043 | hCoV-19/Wales/PHWC-4A1780/2020\|EPI_ISL_743722\|2020 | ···················· | ···················· | ···················· | ···················· | ···················· | ···················· |
| Z | 5044 | hCoV-19/Wales/PHWC-4A17CC/2020\|EPI_ISL_743726\|2020 | ···················· | ··········A········· | ···················· | ···················· | ···················· | ···················· |
| Z | 5045 | hCoV-19/Wales/PHWC-4A17DB/2020\|EPI_ISL_743727\|2020 | ···················· | ··········A········· | ···················· | ···················· | ···················· | ···················· |
| Z | 5048 | hCoV-19/Wales/PHWC-4A1805/2020\|EPI_ISL_743730\|2020 | ···················· | ···················· | ···················· | ···················· | ···················· | ···················· |
| Z | 5049 | hCoV-19/Wales/PHWC-4A1814/2020\|EPI_ISL_743731\|2020 | ···················· | ···················· | ···················· | ···················· | ···················· | ···················· |
| Z | 5050 | hCoV-19/Wales/PHWC-4A1823/2020\|EPI_ISL_743732\|2020 | ···················· | ···················· | ···················· | ···················· | ···················· | ···················· |
| Z | 5051 | hCoV-19/Wales/PHWC-4A1832/2020\|EPI_ISL_743733\|2020 | ···················· | ···················· | ···················· | ···················· | ···················· | ···················· |
| Z | 5052 | hCoV-19/Wales/PHWC-4A1841/2020\|EPI_ISL_743734\|2020 | ···················· | ···················· | ···················· | ···················· | ···················· | ···················· |
| Z | 5053 | hCoV-19/Wales/PHWC-4A1850/2020\|EPI_ISL_743735\|2020 | ···················· | ···················· | ···················· | ···················· | ···················· | ···················· |
| Z | 5054 | hCoV-19/Wales/PHWC-4A187E/2020\|EPI_ISL_743736\|2020 | ···················· | ···················· | ···················· | ···················· | ···················· | ···················· |
| Z | 5055 | hCoV-19/Wales/PHWC-4A188D/2020\|EPI_ISL_743737\|2020 | ···················· | ···················· | ···················· | ···················· | ···················· | ···················· |
| Z | 5057 | hCoV-19/Wales/PHWC-4A18C9/2020\|EPI_ISL_743741\|2020 | ···················· | ···················· | ···················· | ···················· | ···················· | ···················· |
| Z | 5058 | hCoV-19/Wales/PHWC-4A18D8/2020\|EPI_ISL_743742\|2020 | ···················· | ···················· | ···················· | ···················· | ···················· | ···················· |

| | | | | | | | | |
|---|---|---|---|---|---|---|---|---|
| Z | 5059 hCoV-19/Wales/PHWC-4A18E7/2020\|EPI_ISL_743743\|2020 | ···················· | ···················· | ···················· | ···················· | ···················· | ···················· | |
| Z | 5060 hCoV-19/Wales/PHWC-4A1911/2020\|EPI_ISL_743746\|2020 | ···················· | ···················· | ···················· | ···················· | NNN················· | ···················· | 1 |
| Z | 5061 hCoV-19/Wales/PHWC-4A1920/2020\|EPI_ISL_743747\|2020 | ···················· | ··········A········· | ···················· | ···················· | ···················· | ···················· | |
| Z | 5062 hCoV-19/Wales/PHWC-4A193F/2020\|EPI_ISL_743748\|2020 | ···················· | ···················· | ···················· | ···················· | ···················· | ···················· | |
| Z | 5063 hCoV-19/Wales/PHWC-4A194E/2020\|EPI_ISL_743749\|2020 | ···················· | ···················· | ···················· | ···················· | ···················· | ···················· | |
| Z | 5064 hCoV-19/Wales/PHWC-4A196C/2020\|EPI_ISL_743751\|2020 | ···················· | ···················· | ···················· | ···················· | ···················· | ···················· | |
| Z | 5066 hCoV-19/Wales/PHWC-4A1999/2020\|EPI_ISL_743754\|2020 | ···················· | ···················· | ···················· | ···················· | ···················· | ···················· | |
| Z | 5067 hCoV-19/Wales/PHWC-4A19A8/2020\|EPI_ISL_743755\|2020 | ···················· | ···················· | ···················· | ···················· | ···················· | ···················· | |
| Z | 5068 hCoV-19/Wales/PHWC-4A19B7/2020\|EPI_ISL_743756\|2020 | ···················· | ···················· | ···················· | ···················· | ···················· | ···················· | |
| Z | 5069 hCoV-19/Wales/PHWC-4A19C6/2020\|EPI_ISL_743757\|2020 | ···················· | ···················· | ···················· | ···················· | ···················· | ···················· | |
| Z | 5071 hCoV-19/Wales/PHWC-4A19E4/2020\|EPI_ISL_743759\|2020 | ···················· | ···················· | ···················· | ···················· | ···················· | ···················· | |
| Z | 5072 hCoV-19/Wales/PHWC-4A19F3/2020\|EPI_ISL_743760\|2020 | ···················· | ···················· | ···················· | ···················· | ···················· | ···················· | |
| Z | 5073 hCoV-19/Wales/PHWC-4A1A0F/2020\|EPI_ISL_743761\|2020 | ···················· | ···················· | ···················· | ···················· | ···················· | ···················· | |
| Z | 5074 hCoV-19/Wales/PHWC-4A1A1E/2020\|EPI_ISL_743762\|2020 | ···················· | ···················· | ···················· | ···················· | ···················· | ···················· | |
| Z | 5075 hCoV-19/Wales/PHWC-4A1A2D/2020\|EPI_ISL_743763\|2020 | ···················· | ···················· | ···················· | ···················· | ···················· | ···················· | |
| Z | 5076 hCoV-19/Wales/PHWC-4A1A4B/2020\|EPI_ISL_743765\|2020 | ···················· | ···················· | ···················· | ···················· | ···················· | ···················· | |
| Z | 5077 hCoV-19/Wales/PHWC-4A1A5A/2020\|EPI_ISL_743766\|2020 | ···················· | ···················· | ···················· | ···················· | ···················· | ···················· | |
| Z | 5078 hCoV-19/Wales/PHWC-4A1A69/2020\|EPI_ISL_743767\|2020 | ···················· | ···················· | ···················· | ···················· | ···················· | ···················· | |
| Z | 5079 hCoV-19/Wales/PHWC-4A1A87/2020\|EPI_ISL_743769\|2020 | ···················· | ···················· | ···················· | ···················· | ···················· | ···················· | |
| Z | 5080 hCoV-19/Wales/PHWC-4A1AA5/2020\|EPI_ISL_743771\|2020 | ···················· | ···················· | ···················· | ···················· | ···················· | ···················· | |
| Z | 5082 hCoV-19/Wales/PHWC-4A1AD2/2020\|EPI_ISL_743774\|2020 | ···················· | ···················· | ···················· | ···················· | ···················· | N··················· | |
| Z | 5083 hCoV-19/Wales/PHWC-4A1AE1/2020\|EPI_ISL_743775\|2020 | ···················· | ···················· | ···················· | ···················· | ···················· | ···················· | |
| Z | 5084 hCoV-19/Wales/PHWC-4A1B0C/2020\|EPI_ISL_743777\|2020 | ···················· | ···················· | ···················· | ···················· | ···················· | ···················· | |
| Z | 5086 hCoV-19/Wales/PHWC-4A1B2A/2020\|EPI_ISL_743779\|2020 | ···················· | ···················· | ···················· | ···················· | ···················· | ···················· | |
| Z | 5087 hCoV-19/Wales/PHWC-4A1B39/2020\|EPI_ISL_743780\|2020 | ···················· | ···················· | ···················· | ···················· | ···················· | ···················· | |
| Z | 5088 hCoV-19/Wales/PHWC-4A1B48/2020\|EPI_ISL_743781\|2020 | ···················· | ···················· | ···················· | ···················· | ···················· | ···················· | |
| Z | 5089 hCoV-19/Wales/PHWC-4A1B57/2020\|EPI_ISL_743782\|2020 | ···················· | ···················· | ···················· | ···················· | ···················· | ···················· | |
| Z | 5091 hCoV-19/Wales/PHWC-4A1B84/2020\|EPI_ISL_743785\|2020 | ···················· | ···················· | ···················· | ···················· | ···················· | ···················· | |
| Z | 5092 hCoV-19/Wales/PHWC-4A1BA2/2020\|EPI_ISL_743786\|2020 | ···················· | ···················· | ···················· | ···················· | ···················· | ···················· | |
| Z | 5094 hCoV-19/Wales/PHWC-4A1BEE/2020\|EPI_ISL_743788\|2020 | ···················· | ···················· | ···················· | ···················· | ···················· | ···················· | |
| Z | 5095 hCoV-19/Wales/PHWC-4A1BFD/2020\|EPI_ISL_743789\|2020 | ···················· | ···················· | ···················· | ···················· | T··················· | ···················· | |
| Z | 5097 hCoV-19/Wales/PHWC-4A1C18/2020\|EPI_ISL_743791\|2020 | ···················· | ···················· | ···················· | ···················· | ···················· | ···················· | |
| Z | 5098 hCoV-19/Wales/PHWC-4A1C27/2020\|EPI_ISL_743792\|2020 | ···················· | ···················· | ···················· | ···················· | ···················· | ···················· | |
| Z | 5099 hCoV-19/Wales/PHWC-4A1C45/2020\|EPI_ISL_743794\|2020 | ···················· | ···················· | ···················· | ···················· | ···················· | ···················· | |
| Z | 5100 hCoV-19/Wales/PHWC-4A1C54/2020\|EPI_ISL_743795\|2020 | ···················· | ···················· | ···················· | ···················· | ···················· | ···················· | |
| Z | 5103 hCoV-19/Wales/PHWC-4A1C90/2020\|EPI_ISL_743799\|2020 | ···················· | ···················· | ···················· | ···················· | ···················· | ···················· | |
| Z | 5104 hCoV-19/USA/UT-UPHL-2012521294/2020\|EPI_ISL_746000 | ···················· | ···················· | ···················· | ···················· | ···················· | ···················· | |
| Z | 5105 hCoV-19/USA/UT-UPHL-2012780542/2020\|EPI_ISL_746001 | ···················· | ···················· | ···················· | ···················· | ···················· | ···················· | |
| Z | 5106 hCoV-19/USA/UT-UPHL-2012968336/2020\|EPI_ISL_746002 | ···················· | ···················· | ···················· | ···················· | ···················· | ···················· | |
| Z | 5107 hCoV-19/USA/UT-UPHL-2012665767/2020\|EPI_ISL_746003 | ···················· | ···················· | ···················· | ···················· | ···················· | ···················· | |
| Z | 5108 hCoV-19/USA/UT-UPHL-2012930826/2020\|EPI_ISL_746004 | ···················· | ···················· | ···················· | ···················· | ···················· | ···················· | |
| Z | 5109 hCoV-19/USA/UT-UPHL-2012347682/2020\|EPI_ISL_746005 | ···················· | ·A·················· | ···················· | ···················· | ···················· | ···················· | |
| Z | 5110 hCoV-19/USA/UT-UPHL-2012507906/2020\|EPI_ISL_746006 | ···················· | ···················· | ···················· | ···················· | ···················· | ···················· | |
| Z | 5111 hCoV-19/USA/UT-UPHL-2012491096/2020\|EPI_ISL_746007 | ···················· | ···················· | ···················· | ···················· | ···················· | ···················· | |
| Z | 5112 hCoV-19/USA/UT-UPHL-2012006545/2020\|EPI_ISL_746008 | ···················· | ···················· | ···················· | ···················· | ···················· | ···················· | |
| Z | 5113 hCoV-19/USA/UT-UPHL-2012543435/2020\|EPI_ISL_746009 | ···················· | ···················· | ···················· | ···················· | ···················· | ···················· | |
| Z | 5114 hCoV-19/USA/UT-UPHL-2012618881/2020\|EPI_ISL_746040 | ···················· | ···················· | ···················· | ···················· | ···················· | ···················· | |
| Z | 5115 hCoV-19/USA/UT-UPHL-2012566629/2020\|EPI_ISL_746041 | ···················· | ···················· | ···················· | ···················· | ···················· | ···················· | |
| Z | 5116 hCoV-19/USA/UT-UPHL-2012642882/2020\|EPI_ISL_746042 | ···················· | ···················· | ···················· | ···················· | ···················· | ···················· | |
| Z | 5117 hCoV-19/USA/UT-UPHL-2012427205/2020\|EPI_ISL_746046 | ···················· | ···················· | ···················· | ···················· | ···················· | ···················· | |
| Z | 5118 hCoV-19/USA/UT-UPHL-2012392041/2020\|EPI_ISL_746047 | ···················· | ···················· | ···················· | ···················· | ···················· | ···················· | |
| Z | 5119 hCoV-19/USA/UT-UPHL-2012392157/2020\|EPI_ISL_746048 | ···················· | ···················· | ···················· | ···················· | ···················· | ···················· | |
| Z | 5120 hCoV-19/USA/UT-UPHL-2012859839/2020\|EPI_ISL_746049 | ···················· | ···················· | ···················· | ···················· | ···················· | ···················· | |
| Z | 5121 hCoV-19/England/LOND-1266D57/2020\|EPI_ISL_741204\|2 | ···················· | ···················· | ···················· | ···················· | ···················· | ···················· | |
| Z | 5122 hCoV-19/England/LOND-1266E09/2020\|EPI_ISL_741215\|2 | ···················· | ···················· | ···················· | ···················· | ···················· | ···················· | |
| Z | 5123 hCoV-19/England/LOND-1266E27/2020\|EPI_ISL_741217\|2 | ···················· | ···················· | ···················· | ···················· | ···················· | ···················· | |
| Z | 5124 hCoV-19/England/LOND-1266E63/2020\|EPI_ISL_741221\|2 | ···················· | ···················· | ···················· | ···················· | ······T············· | ···················· | |
| Z | 5125 hCoV-19/England/LOND-1266E72/2020\|EPI_ISL_741222\|2 | ···················· | ···················· | ···················· | ···················· | ···················· | ···················· | |
| Z | 5126 hCoV-19/England/LOND-1266E81/2020\|EPI_ISL_741223\|2 | ···················· | ···················· | ···················· | ···················· | ···················· | ···················· | |
| Z | 5127 hCoV-19/England/LOND-1266EAF/2020\|EPI_ISL_741225\|2 | ···················· | ···················· | ···················· | ···················· | ···················· | ···················· | |
| Z | 5128 hCoV-19/England/LOND-1266EBE/2020\|EPI_ISL_741226\|2 | ···················· | ···················· | ···················· | ···················· | ······T············· | ···················· | |
| Z | 5129 hCoV-19/England/LOND-1266FBB/2020\|EPI_ISL_741238\|2 | ···················· | ···················· | ···················· | ···················· | ······T············· | ···················· | |
| Z | 5130 hCoV-19/England/LOND-126705D/2020\|EPI_ISL_741246\|2 | ···················· | ···················· | ···················· | ···················· | ···················· | ···················· | |
| Z | 5131 hCoV-19/England/LOND-1267099/2020\|EPI_ISL_741250\|2 | ···················· | ···················· | ···················· | ···················· | ···················· | ···················· | |
| Z | 5132 hCoV-19/England/LOND-126721B/2020\|EPI_ISL_741260\|2 | ···················· | ···················· | ···················· | ···················· | ···················· | ···················· | |
| Z | 5133 hCoV-19/England/LOND-1267275/2020\|EPI_ISL_741264\|2 | ···················· | ···················· | ···················· | ···················· | ···················· | ···················· | |
| Z | 5134 hCoV-19/England/LOND-1267309/2020\|EPI_ISL_741270\|2 | ···················· | ···················· | ···················· | ···················· | ···················· | ···················· | |
| Z | 5135 hCoV-19/England/LOND-1267354/2020\|EPI_ISL_741275\|2 | ···················· | ···················· | ···················· | ···················· | ···················· | ···················· | |
| Z | 5136 hCoV-19/England/LOND-1267363/2020\|EPI_ISL_741276\|2 | ···················· | ···················· | ···················· | ···················· | ···················· | ···················· | |
| Z | 5137 hCoV-19/England/LOND-1267390/2020\|EPI_ISL_741279\|2 | ···················· | ···················· | ···················· | ···················· | ······T············· | ···················· | |
| Z | 5138 hCoV-19/England/LOND-1267406/2020\|EPI_ISL_741285\|2 | ···················· | ···················· | ···················· | ···················· | ···················· | ···················· | |
| Z | 5139 hCoV-19/England/LOND-1267460/2020\|EPI_ISL_741291\|2 | ···················· | ···················· | ···················· | ···················· | ···················· | ···················· | |
| Z | 5140 hCoV-19/England/LOND-12674AC/2020\|EPI_ISL_741295\|2 | ···················· | ···················· | ···················· | ···················· | ···················· | ···················· | |
| Z | 5141 hCoV-19/England/LOND-12674CA/2020\|EPI_ISL_741297\|2 | ···················· | ·A·················· | ···················· | ···················· | ···················· | ···A················ | |
| Z | 5142 hCoV-19/England/LOND-12674D9/2020\|EPI_ISL_741298\|2 | ···················· | ···················· | ···················· | ···················· | ···················· | ···················· | |
| Z | 5143 hCoV-19/Wales/PHWC-49C07E/2020\|EPI_ISL_742501\|2020 | ···················· | ···················· | ···················· | ···················· | ···················· | ···················· | |
| Z | 5145 hCoV-19/Wales/PHWC-49C0AB/2020\|EPI_ISL_742504\|2020 | ···················· | ···················· | ···················· | ···················· | ···················· | ···················· | |
| Z | 5149 hCoV-19/Wales/PHWC-49C102/2020\|EPI_ISL_742509\|2020 | ···················· | ···················· | ···················· | ···················· | ···················· | ···················· | |
| Z | 5153 hCoV-19/Wales/PHWC-49C15D/2020\|EPI_ISL_742514\|2020 | ···················· | ···················· | ···················· | ···················· | ···················· | ···················· | |
| Z | 5154 hCoV-19/Wales/PHWC-49C17B/2020\|EPI_ISL_742516\|2020 | ···················· | ···················· | ···················· | ···················· | ···················· | ···················· | |

| | | | | | | | | |
|---|---|---|---|---|---|---|---|---|
| Z | 5157 | hCoV-19/Wales/PHWC-49C1A8/2020\|EPI_ISL_742519\|2020 | ···················· | ···················· | ···················· | ···················· | ···················· | ···················· |
| Z | 5158 | hCoV-19/Wales/PHWC-49C1B7/2020\|EPI_ISL_742520\|2020 | ···················· | ···················· | ···················· | ···················· | ···················· | ···················· |
| Z | 5159 | hCoV-19/Wales/PHWC-49C1C6/2020\|EPI_ISL_742521\|2020 | ···················· | ···················· | ···················· | ···················· | ···················· | ···················· |
| Z | 5161 | hCoV-19/Wales/PHWC-49C1F3/2020\|EPI_ISL_742524\|2020 | ···················· | ···················· | ···················· | ···················· | ···················· | ···················· |
| Z | 5162 | hCoV-19/Wales/PHWC-49C21E/2020\|EPI_ISL_742526\|2020 | ···················· | ···················· | ···················· | ···················· | ···················· | ···················· |
| Z | 5166 | hCoV-19/Wales/PHWC-49C287/2020\|EPI_ISL_742533\|2020 | ···················· | ···················· | ···················· | ···················· | ···················· | ···················· |
| Z | 5168 | hCoV-19/Wales/PHWC-49C2B4/2020\|EPI_ISL_742536\|2020 | ···················· | ···················· | ···················· | ···················· | ···················· | ···················· |
| Z | 5170 | hCoV-19/Wales/PHWC-49C2E1/2020\|EPI_ISL_742539\|2020 | ···················· | ···················· | ···················· | ···················· | ···················· | ···················· |
| Z | 5172 | hCoV-19/Wales/PHWC-49C30C/2020\|EPI_ISL_742541\|2020 | ···················· | ···················· | ···················· | ···················· | ···················· | ···················· |
| Z | 5173 | hCoV-19/Wales/PHWC-49C31B/2020\|EPI_ISL_742542\|2020 | ···················· | ···················· | ···················· | ···················· | ···················· | ···················· |
| Z | 5174 | hCoV-19/Wales/PHWC-49C32A/2020\|EPI_ISL_742543\|2020 | ···················· | ···················· | ···················· | ···················· | ···················· | ···················· |
| Z | 5175 | hCoV-19/Wales/PHWC-49C339/2020\|EPI_ISL_742544\|2020 | ···················· | ···················· | ···················· | ···················· | ···················· | ···················· |
| Z | 5176 | hCoV-19/Wales/PHWC-49C357/2020\|EPI_ISL_742545\|2020 | ···················· | ···················· | ···················· | ···················· | ···················· | ···················· |
| Z | 5177 | hCoV-19/Wales/PHWC-49C366/2020\|EPI_ISL_742546\|2020 | ···················· | ···················· | ···················· | ···················· | ···················· | ···················· |
| Z | 5178 | hCoV-19/Wales/PHWC-49C375/2020\|EPI_ISL_742547\|2020 | ···················· | ···················· | ···················· | ···················· | ···················· | ···················· |
| Z | 5179 | hCoV-19/Wales/PHWC-49C384/2020\|EPI_ISL_742548\|2020 | ···················· | ···················· | ···················· | ···················· | ···················· | ···················· |
| Z | 5180 | hCoV-19/Wales/PHWC-49C3B1/2020\|EPI_ISL_742550\|2020 | ···················· | ···················· | ···················· | ···················· | ···················· | ···················· |
| Z | 5181 | hCoV-19/Wales/PHWC-49C3C0/2020\|EPI_ISL_742551\|2020 | ···················· | ···················· | ···················· | ···················· | ···················· | ···················· |
| Z | 5182 | hCoV-19/Wales/PHWC-49C3DF/2020\|EPI_ISL_742552\|2020 | ···················· | ···················· | ···················· | ···················· | ···················· | ···················· |
| Z | 5183 | hCoV-19/Wales/PHWC-49C3FD/2020\|EPI_ISL_742554\|2020 | ···················· | ···················· | ···················· | ···················· | ···················· | ···················· |
| Z | 5184 | hCoV-19/Wales/PHWC-49C418/2020\|EPI_ISL_742555\|2020 | ···················· | ···················· | ···················· | ···················· | ···················· | ···················· |
| Z | 5185 | hCoV-19/Wales/PHWC-49C427/2020\|EPI_ISL_742556\|2020 | ···················· | ···················· | ···················· | ···················· | ···················· | ···················· |
| Z | 5186 | hCoV-19/Wales/PHWC-49C436/2020\|EPI_ISL_742557\|2020 | ···················· | ···················· | ···················· | ···················· | ···················· | ···················· |
| Z | 5187 | hCoV-19/Wales/PHWC-49C445/2020\|EPI_ISL_742558\|2020 | ···················· | ···················· | ···················· | ···················· | ···················· | ···················· |
| Z | 5188 | hCoV-19/Wales/PHWC-49C472/2020\|EPI_ISL_742559\|2020 | ···················· | ···················· | ···················· | ···················· | ···················· | ···················· |
| Z | 5189 | hCoV-19/Wales/PHWC-49C481/2020\|EPI_ISL_742560\|2020 | ···················· | ···················· | ···················· | ···················· | ···················· | ···················· |
| Z | 5190 | hCoV-19/Wales/PHWC-49C4AF/2020\|EPI_ISL_742561\|2020 | ···················· | ···················· | ···················· | ···················· | ···················· | ···················· |
| Z | 5191 | hCoV-19/Wales/PHWC-49C4BE/2020\|EPI_ISL_742562\|2020 | ···················· | ···················· | ···················· | ···················· | ···················· | ···················· |
| Z | 5193 | hCoV-19/Wales/PHWC-49C4FA/2020\|EPI_ISL_742565\|2020 | ···················· | ···················· | ···················· | ···················· | ···················· | ···················· |
| Z | 5194 | hCoV-19/Wales/PHWC-49C515/2020\|EPI_ISL_742566\|2020 | ···················· | ···················· | ···················· | ···················· | ···················· | ···················· |
| Z | 5195 | hCoV-19/Wales/PHWC-49C533/2020\|EPI_ISL_742568\|2020 | ···················· | ···················· | ···················· | ···················· | ···················· | ···················· |
| Z | 5196 | hCoV-19/Wales/PHWC-49C542/2020\|EPI_ISL_742569\|2020 | ···················· | ···················· | ···················· | ···················· | ···················· | ···················· |
| Z | 5197 | hCoV-19/Wales/PHWC-49C560/2020\|EPI_ISL_742571\|2020 | ···················· | ···················· | ···················· | ···················· | ···················· | ···················· |
| Z | 5198 | hCoV-19/Wales/PHWC-49C58E/2020\|EPI_ISL_742573\|2020 | ···················· | ···················· | ···················· | ···················· | ···················· | ···················· |
| Z | 5199 | hCoV-19/Wales/PHWC-49C5AC/2020\|EPI_ISL_742575\|2020 | ···················· | ···················· | ···················· | ···················· | ···················· | ···················· |
| Z | 5200 | hCoV-19/Wales/PHWC-49C5BB/2020\|EPI_ISL_742576\|2020 | ···················· | ···················· | ···················· | ···················· | ···················· | ···················· |
| Z | 5201 | hCoV-19/Wales/PHWC-49C5D9/2020\|EPI_ISL_742578\|2020 | ···················· | ···················· | ···················· | ···················· | ···················· | ···················· |
| Z | 5202 | hCoV-19/Wales/PHWC-49C5E8/2020\|EPI_ISL_742579\|2020 | ···················· | ···················· | ···················· | ···················· | ···················· | ···················· |
| Z | 5203 | hCoV-19/Wales/PHWC-49C5F7/2020\|EPI_ISL_742580\|2020 | ···················· | ···················· | ···················· | ···················· | ···················· | ···················· |
| Z | 5204 | hCoV-19/Wales/PHWC-49C603/2020\|EPI_ISL_742581\|2020 | ···················· | ···················· | ···················· | ···················· | ···················· | ···················· |
| Z | 5205 | hCoV-19/Wales/PHWC-49C612/2020\|EPI_ISL_742582\|2020 | ···················· | ···················· | ···················· | ···················· | ···················· | ···················· |
| Z | 5206 | hCoV-19/Wales/PHWC-49C621/2020\|EPI_ISL_742583\|2020 | ···················· | ···················· | ···················· | ···················· | ···················· | ···················· |
| Z | 5207 | hCoV-19/Wales/PHWC-49C630/2020\|EPI_ISL_742584\|2020 | ···················· | ···················· | ···················· | ···················· | ···················· | ···················· |
| Z | 5208 | hCoV-19/Wales/PHWC-49C64F/2020\|EPI_ISL_742585\|2020 | ···················· | ···················· | ···················· | ···················· | ···················· | ···················· |
| Z | 5210 | hCoV-19/Wales/PHWC-49C66D/2020\|EPI_ISL_742587\|2020 | ···················· | ···················· | ···················· | ···················· | ···················· | ···················· |
| Z | 5211 | hCoV-19/Wales/PHWC-49C68B/2020\|EPI_ISL_742588\|2020 | ···················· | ···················· | ···················· | ···················· | ···················· | ···················· |
| Z | 5212 | hCoV-19/Wales/PHWC-49C69A/2020\|EPI_ISL_742589\|2020 | ···················· | ···················· | ···················· | ···················· | ···················· | ···················· |
| Z | 5213 | hCoV-19/Wales/PHWC-49C6A9/2020\|EPI_ISL_742590\|2020 | ···················· | ···················· | ···················· | ···················· | ···················· | ···················· |
| Z | 5214 | hCoV-19/Wales/PHWC-49C6D6/2020\|EPI_ISL_742592\|2020 | ···················· | ···················· | ···················· | ···················· | ···················· | ···················· |
| Z | 5215 | hCoV-19/Wales/PHWC-49C6F4/2020\|EPI_ISL_742594\|2020 | ···················· | ···················· | ···················· | ···················· | ···················· | ···················· |
| Z | 5216 | hCoV-19/Wales/PHWC-49C73D/2020\|EPI_ISL_742596\|2020 | ···················· | ···················· | ···················· | ···················· | ···················· | ···················· |
| Z | 5217 | hCoV-19/Wales/PHWC-49C76A/2020\|EPI_ISL_742597\|2020 | ···················· | ···················· | ···················· | ···················· | ···················· | ···················· |
| Z | 5218 | hCoV-19/Wales/PHWC-49C779/2020\|EPI_ISL_742598\|2020 | ···················· | ···················· | ···················· | ···················· | ···A················ | ···················· |
| Z | 5219 | hCoV-19/Wales/PHWC-49C788/2020\|EPI_ISL_742599\|2020 | ···················· | ···················· | ···················· | ···················· | ···················· | ···················· |
| Z | 5220 | hCoV-19/Wales/PHWC-4A1CAF/2020\|EPI_ISL_743800\|2020 | ···················· | ···················· | ···················· | ···················· | ···················· | ···················· |
| Z | 5222 | hCoV-19/Wales/PHWC-4A1D06/2020\|EPI_ISL_743805\|2020 | ···················· | ···················· | ···················· | ···················· | ···················· | ···················· |
| Z | 5224 | hCoV-19/Wales/PHWC-4A1D51/2020\|EPI_ISL_743808\|2020 | ···················· | ···················· | ···················· | ···················· | ···················· | ···················· |
| Z | 5225 | hCoV-19/Wales/PHWC-4A1D8E/2020\|EPI_ISL_743811\|2020 | ···················· | ···················· | ···················· | ···················· | ···················· | ···················· |
| Z | 5226 | hCoV-19/Wales/PHWC-4A1D9D/2020\|EPI_ISL_743812\|2020 | ···················· | ···················· | ···················· | ···················· | ···················· | ···················· |
| Z | 5227 | hCoV-19/Wales/PHWC-4A1DCA/2020\|EPI_ISL_743815\|2020 | ···················· | ···················· | ···················· | ···················· | ···················· | ···················· |
| Z | 5228 | hCoV-19/Wales/PHWC-4A1DD9/2020\|EPI_ISL_743816\|2020 | ···················· | ···················· | ···················· | ···················· | ···················· | ···················· |
| Z | 5229 | hCoV-19/Wales/PHWC-4A1DE8/2020\|EPI_ISL_743817\|2020 | ···················· | ···················· | ···················· | ···················· | ···················· | ···················· |
| Z | 5230 | hCoV-19/Wales/PHWC-4A1DF7/2020\|EPI_ISL_743818\|2020 | ···················· | ···················· | ···················· | ···················· | ···················· | ···················· |
| Z | 5231 | hCoV-19/Wales/PHWC-4A1E03/2020\|EPI_ISL_743819\|2020 | ···················· | ···················· | ···················· | ···················· | ···················· | ···················· |
| Z | 5232 | hCoV-19/Wales/PHWC-4A1E4F/2020\|EPI_ISL_743822\|2020 | ···················· | ···················· | ···················· | ···················· | ···················· | ···················· |
| Z | 5233 | hCoV-19/Wales/PHWC-4A1E5E/2020\|EPI_ISL_743823\|2020 | ···················· | ···················· | ···················· | ···················· | ···················· | ···················· |
| Z | 5234 | hCoV-19/Wales/PHWC-4A1E6D/2020\|EPI_ISL_743824\|2020 | ···················· | ···················· | ···················· | ···················· | ···················· | ···················· |
| Z | 5236 | hCoV-19/Wales/PHWC-4A1E9A/2020\|EPI_ISL_743826\|2020 | ···················· | ···················· | ···················· | ···················· | ···················· | ···················· |
| Z | 5237 | hCoV-19/Wales/PHWC-4A1EA9/2020\|EPI_ISL_743827\|2020 | ···················· | ···················· | ···················· | ···················· | ···················· | ···················· |
| Z | 5238 | hCoV-19/Wales/PHWC-4A1EC7/2020\|EPI_ISL_743828\|2020 | ···················· | ···················· | ···················· | ···················· | ···················· | ···················· |
| Z | 5239 | hCoV-19/Wales/PHWC-4A1EE5/2020\|EPI_ISL_743829\|2020 | ···················· | ···················· | ···················· | ···················· | ···················· | ···················· |
| Z | 5240 | hCoV-19/Wales/PHWC-4A1EF4/2020\|EPI_ISL_743830\|2020 | ···················· | ···················· | ···················· | ···················· | ···················· | ···················· |
| Z | 5241 | hCoV-19/Wales/PHWC-4A1F00/2020\|EPI_ISL_743831\|2020 | ···················· | ···················· | ···················· | ···················· | ···················· | ···················· |
| Z | 5242 | hCoV-19/Wales/PHWC-4A1F5B/2020\|EPI_ISL_743834\|2020 | ···················· | ···················· | ···················· | ···················· | ···················· | ···················· |
| Z | 5243 | hCoV-19/Wales/PHWC-4A1F6A/2020\|EPI_ISL_743835\|2020 | ···················· | ···················· | ···················· | ···················· | ···················· | ···················· |
| Z | 5244 | hCoV-19/Wales/PHWC-4A1F88/2020\|EPI_ISL_743837\|2020 | ···················· | ···················· | ···················· | ···················· | ···················· | ···················· |
| Z | 5245 | hCoV-19/Wales/PHWC-4A1F97/2020\|EPI_ISL_743838\|2020 | ···················· | ···················· | ···················· | ···················· | ···················· | ···················· |
| Z | 5247 | hCoV-19/Wales/PHWC-4A1FE2/2020\|EPI_ISL_743841\|2020 | ···················· | ···················· | ···················· | ···················· | ···················· | ···················· |
| Z | 5255 | hCoV-19/Wales/PHWC-4A2172/2020\|EPI_ISL_743853\|2020 | ···················· | ···················· | ···················· | ···················· | ···················· | ···················· |
| Z | 5256 | hCoV-19/Wales/PHWC-4A2181/2020\|EPI_ISL_743854\|2020 | ···················· | ···················· | ···················· | ···················· | ···················· | ···················· |

| | | | | | | | |
|---|---|---|---|---|---|---|---|
| Z | 5257 hCoV-19/Wales/PHWC-4A2190/2020\|EPI_ISL_743855\|2020 | .................... | .................... | .................... | .................... | .................... | .................... |
| Z | 5258 hCoV-19/Wales/PHWC-4A21AF/2020\|EPI_ISL_743856\|2020 | .................... | .................... | .................... | .................... | .................... | .................... |
| Z | 5259 hCoV-19/Wales/PHWC-4A21BE/2020\|EPI_ISL_743857\|2020 | .................... | .................... | .................... | .................... | .................... | .................... |
| Z | 5260 hCoV-19/Wales/PHWC-4A21CD/2020\|EPI_ISL_743858\|2020 | .................... | .................... | .................... | .................... | .................... | .................... |
| Z | 5261 hCoV-19/Wales/PHWC-4A21DC/2020\|EPI_ISL_743859\|2020 | .................... | .................... | .................... | .................... | .................... | .................... |
| Z | 5262 hCoV-19/Wales/PHWC-4A21FA/2020\|EPI_ISL_743861\|2020 | .................... | .................... | .................... | .................... | .................... | .................... |
| Z | 5264 hCoV-19/Wales/PHWC-4A2251/2020\|EPI_ISL_743863\|2020 | .................... | .................... | .................... | .................... | .................... | .................... |
| Z | 5265 hCoV-19/Wales/PHWC-4A2260/2020\|EPI_ISL_743864\|2020 | .................... | .................... | .................... | .................... | .................... | .................... |
| Z | 5266 hCoV-19/Wales/PHWC-4A228E/2020\|EPI_ISL_743866\|2020 | .................... | .................... | .................... | .................... | .................... | .................... |
| Z | 5268 hCoV-19/Wales/PHWC-4A22AC/2020\|EPI_ISL_743868\|2020 | .................... | .................... | .................... | .................... | .................... | .................... |
| Z | 5269 hCoV-19/Wales/PHWC-4A22CA/2020\|EPI_ISL_743870\|2020 | .................... | .................... | .................... | .................... | .................... | .................... |
| Z | 5270 hCoV-19/Wales/PHWC-4A22D9/2020\|EPI_ISL_743871\|2020 | .................... | .................... | .................... | .................... | .................... | .................... |
| Z | 5271 hCoV-19/Wales/PHWC-4A2330/2020\|EPI_ISL_743874\|2020 | .................... | .................... | .................... | .................... | .................... | .................... |
| Z | 5272 hCoV-19/Wales/PHWC-4A234F/2020\|EPI_ISL_743875\|2020 | .................... | .................... | .................... | .................... | .................... | .................... |
| Z | 5273 hCoV-19/Wales/PHWC-4A235E/2020\|EPI_ISL_743876\|2020 | .................... | .................... | .................... | .................... | .................... | .................... |
| Z | 5274 hCoV-19/Wales/PHWC-4A236D/2020\|EPI_ISL_743877\|2020 | .................... | .................... | .................... | .................... | .................... | .................... |
| Z | 5275 hCoV-19/Wales/PHWC-4A237C/2020\|EPI_ISL_743878\|2020 | .................... | .................... | .................... | .................... | .................... | .................... |
| Z | 5276 hCoV-19/Wales/PHWC-4A238B/2020\|EPI_ISL_743879\|2020 | .................... | .................... | .................... | .................... | .................... | .................... |
| Z | 5277 hCoV-19/Wales/PHWC-4A23A9/2020\|EPI_ISL_743880\|2020 | .................... | .................... | .................... | .................... | .................... | .................... |
| Z | 5278 hCoV-19/Wales/PHWC-4A23B8/2020\|EPI_ISL_743881\|2020 | .................... | .................... | .................... | .................... | .................... | .................... |
| Z | 5280 hCoV-19/Wales/PHWC-4A23D6/2020\|EPI_ISL_743883\|2020 | .................... | .................... | .................... | .................... | .................... | .................... |
| Z | 5281 hCoV-19/Wales/PHWC-4A23E5/2020\|EPI_ISL_743884\|2020 | .................... | .................... | .................... | .................... | .................... | .................... |
| Z | 5282 hCoV-19/Wales/PHWC-4A23F4/2020\|EPI_ISL_743885\|2020 | .................... | .................... | .................... | .................... | .................... | .................... |
| Z | 5283 hCoV-19/Wales/PHWC-4A2400/2020\|EPI_ISL_743886\|2020 | .................... | .................... | .................... | .................... | .................... | .................... |
| Z | 5284 hCoV-19/Wales/PHWC-4A241F/2020\|EPI_ISL_743887\|2020 | .................... | .................... | .................... | .................... | .................... | .................... |
| Z | 5285 hCoV-19/Wales/PHWC-4A242E/2020\|EPI_ISL_743888\|2020 | .................... | .................... | .................... | .................... | .................... | .................... |
| Z | 5286 hCoV-19/Wales/PHWC-4A243D/2020\|EPI_ISL_743889\|2020 | .................... | .................... | .................... | .................... | .................... | .................... |
| Z | 5287 hCoV-19/Wales/PHWC-4A245B/2020\|EPI_ISL_743890\|2020 | .................... | .................... | .................... | .................... | .................... | .................... |
| Z | 5288 hCoV-19/Wales/PHWC-4A2497/2020\|EPI_ISL_743893\|2020 | .................... | .................... | .................... | .................... | .................... | .................... |
| Z | 5289 hCoV-19/Wales/PHWC-4A24A6/2020\|EPI_ISL_743894\|2020 | .................... | .................... | .................... | .................... | .................... | .................... |
| Z | 5290 hCoV-19/Wales/PHWC-4A24C4/2020\|EPI_ISL_743896\|2020 | .................... | .................... | .................... | .................... | .................... | .................... |
| Z | 5291 hCoV-19/Wales/PHWC-4A24D3/2020\|EPI_ISL_743897\|2020 | .................... | .................... | .................... | .................... | .................... | .................... |
| Z | 5292 hCoV-19/Wales/PHWC-4A24E2/2020\|EPI_ISL_743898\|2020 | .................... | .................... | .................... | .................... | .................... | .................... |
| Z | 5293 hCoV-19/Wales/PHWC-4A24F1/2020\|EPI_ISL_743899\|2020 | .................... | .................... | .................... | .................... | .................... | .................... |
| Z | 5294 hCoV-19/USA/UT-UPHL-2012395754/2020\|EPI_ISL_746010 | .................... | .................... | .................... | .................... | .................... | .................... |
| Z | 5295 hCoV-19/USA/UT-UPHL-2012993951/2020\|EPI_ISL_746011 | .................... | .................... | .................... | .................... | .................... | .................... |
| Z | 5296 hCoV-19/USA/UT-UPHL-2012972605/2020\|EPI_ISL_746012 | .................... | .................... | .................... | .................... | .................... | .................... |
| Z | 5297 hCoV-19/USA/UT-UPHL-2012754446/2020\|EPI_ISL_746013 | .................... | .................... | .................... | .................... | .................... | .................... |
| Z | 5298 hCoV-19/USA/UT-UPHL-2012212284/2020\|EPI_ISL_746014 | .................... | .................... | .................... | .................... | .................... | .................... |
| Z | 5299 hCoV-19/USA/UT-UPHL-2012291116/2020\|EPI_ISL_746015 | .................... | .A.................. | .................... | .................... | .................... | .................... |
| Z | 5300 hCoV-19/USA/UT-UPHL-2012705073/2020\|EPI_ISL_746016 | .................... | .A.................. | .................... | .................... | .................... | .................... |
| Z | 5301 hCoV-19/USA/UT-UPHL-2012776828/2020\|EPI_ISL_746017 | .................... | .................... | .................... | .................... | .................... | .................... |
| Z | 5302 hCoV-19/USA/UT-UPHL-2012829129/2020\|EPI_ISL_746018 | .................... | .................... | .................... | .................... | .................... | .................... |
| Z | 5303 hCoV-19/USA/UT-UPHL-2012019907/2020\|EPI_ISL_746019 | .................... | .................... | .................... | .................... | .................... | .................... |
| Z | 5304 hCoV-19/USA/UT-UPHL-2012716871/2020\|EPI_ISL_746020 | .................... | .A.................. | .................... | .................... | .................... | .................... |
| Z | 5305 hCoV-19/USA/UT-UPHL-2012629112/2020\|EPI_ISL_746021 | .................... | .................... | .................... | .................... | .................... | .................... |
| Z | 5306 hCoV-19/USA/UT-UPHL-2012828622/2020\|EPI_ISL_746022 | .................... | .................... | .................... | .................... | .................... | .................... |
| Z | 5307 hCoV-19/USA/UT-UPHL-2012259762/2020\|EPI_ISL_746023 | .................... | .................... | .................... | .................... | .................... | .................... |
| Z | 5308 hCoV-19/USA/UT-UPHL-2012185803/2020\|EPI_ISL_746024 | .................... | .................... | .................... | .................... | .................... | .................... |
| Z | 5309 hCoV-19/USA/UT-UPHL-2012698910/2020\|EPI_ISL_746025 | .................... | .................... | .................... | .................... | .................... | .................... |
| Z | 5310 hCoV-19/USA/UT-UPHL-2012139539/2020\|EPI_ISL_746026 | .................... | .................... | .................... | .................... | .................... | .................... |
| Z | 5311 hCoV-19/USA/UT-UPHL-2012021102/2020\|EPI_ISL_746027 | .................... | .................... | .................... | .................... | .................... | .................... |
| Z | 5312 hCoV-19/USA/UT-UPHL-2012049713/2020\|EPI_ISL_746028 | .................... | .................... | .................... | .................... | .................... | .................... |
| Z | 5313 hCoV-19/USA/UT-UPHL-2012736922/2020\|EPI_ISL_746029 | .................... | .................... | .................... | .................... | .................... | .................... |
| Z | 5314 hCoV-19/USA/UT-UPHL-2012481939/2020\|EPI_ISL_746030 | .................... | .................... | .................... | .................... | .................... | .................... |
| Z | 5315 hCoV-19/USA/UT-UPHL-2012198597/2020\|EPI_ISL_746031 | .................... | .A.................. | .................... | .................... | .................... | .................... |
| Z | 5316 hCoV-19/USA/UT-UPHL-2012289777/2020\|EPI_ISL_746032 | .................... | .A.................. | .................... | .................... | .................... | .................... |
| Z | 5317 hCoV-19/USA/UT-UPHL-2012651246/2020\|EPI_ISL_746033 | .................... | .A.................. | .................... | .................... | .................... | .................... |
| Z | 5318 hCoV-19/USA/UT-UPHL-2012630521/2020\|EPI_ISL_746034 | .................... | .................... | .................... | .................... | .................... | .................... |
| Z | 5319 hCoV-19/USA/UT-UPHL-2012898851/2020\|EPI_ISL_746035 | .................... | .................... | .................... | .................... | .................... | .................... |
| Z | 5320 hCoV-19/USA/UT-UPHL-2012875970/2020\|EPI_ISL_746036 | .................... | .................... | .................... | .................... | .................... | .................... |
| Z | 5321 hCoV-19/USA/UT-UPHL-2012036481/2020\|EPI_ISL_746037 | .................... | .................... | .................... | .................... | .................... | .................... |
| Z | 5322 hCoV-19/USA/UT-UPHL-2012826103/2020\|EPI_ISL_746038 | .................... | .A.................. | .................... | .................... | .................... | .................... |
| Z | 5323 hCoV-19/USA/UT-UPHL-2012967418/2020\|EPI_ISL_746039 | .................... | .................... | .................... | .................... | .................... | .................... |
| Z | 5324 hCoV-19/USA/UT-UPHL-2012096240/2020\|EPI_ISL_746043 | .................... | .................... | .................... | .................... | .................... | .................... |
| Z | 5325 hCoV-19/USA/UT-UPHL-2012835843/2020\|EPI_ISL_746044 | .................... | .................... | .................... | .................... | .................... | .................... |
| Z | 5326 hCoV-19/USA/UT-UPHL-2012362641/2020\|EPI_ISL_746045 | .................... | .................... | .................... | .................... | .................... | .................... |
| Z | 5327 hCoV-19/USA/UT-UPHL-2012081770/2020\|EPI_ISL_746050 | .................... | .A.................. | .................... | .................... | .................... | .................... |
| Z | 5328 hCoV-19/USA/UT-UPHL-2012754321/2020\|EPI_ISL_746051 | .................... | .A.................. | .................... | .................... | .................... | .................... |
| Z | 5329 hCoV-19/USA/UT-UPHL-2012733496/2020\|EPI_ISL_746052 | .................... | .A.................. | .................... | .................... | .................... | .................... |
| Z | 5330 hCoV-19/USA/UT-UPHL-2012965262/2020\|EPI_ISL_746053 | .................... | .A.................. | .................... | .................... | .................... | .................... |
| Z | 5331 hCoV-19/USA/UT-UPHL-2012762917/2020\|EPI_ISL_746054 | .................... | .................... | .................... | .................... | .................... | .................... |
| Z | 5332 hCoV-19/USA/UT-UPHL-2012309299/2020\|EPI_ISL_746055 | .................... | .................... | .................... | .................... | .................... | .................... |
| Z | 5333 hCoV-19/USA/UT-UPHL-2012868464/2020\|EPI_ISL_746056 | .................... | .A.................. | .................... | .................... | .................... | .................... |
| Z | 5334 hCoV-19/USA/UT-UPHL-2012022026/2020\|EPI_ISL_746057 | .................... | .................... | .................... | .................... | .................... | .................... |
| Z | 5335 hCoV-19/USA/UT-UPHL-2012634965/2020\|EPI_ISL_746058 | .................... | .A.................. | .................... | .................... | .................... | .................... |
| Z | 5336 hCoV-19/USA/UT-UPHL-2012303126/2020\|EPI_ISL_746059 | .................... | .................... | .................... | .................... | .................... | .................... |
| Z | 5337 hCoV-19/USA/UT-UPHL-2012666124/2020\|EPI_ISL_746060 | .................... | .................... | .................... | .................... | .................... | .................... |
| Z | 5338 hCoV-19/USA/UT-UPHL-2012361771/2020\|EPI_ISL_746061 | .................... | .................... | .................... | .................... | .................... | .................... |
| Z | 5339 hCoV-19/USA/UT-UPHL-2012196020/2020\|EPI_ISL_746062 | .................... | .................... | .................... | .................... | .................... | .................... |

| | | | | | | | | |
|---|---|---|---|---|---|---|---|---|
| Z | 5340 | hCoV-19/USA/UT-UPHL-2012913551/2020\|EPI_ISL_746063 | ···················· | ···················· | ···················· | ···················· | ···················· | ···················· |
| Z | 5341 | hCoV-19/USA/UT-UPHL-2012916593/2020\|EPI_ISL_746064 | ···················· | ···················· | ···················· | ···················· | ···················· | ···················· |
| Z | 5342 | hCoV-19/USA/UT-UPHL-2012164094/2020\|EPI_ISL_746065 | ···················· | ·A·················· | ···················· | ···················· | ···················· | ···················· |
| Z | 5343 | hCoV-19/USA/UT-UPHL-2012610616/2020\|EPI_ISL_746066 | ···················· | ···················· | ···················· | ···················· | ···················· | ···················· |
| Z | 5344 | hCoV-19/USA/UT-UPHL-2012580181/2020\|EPI_ISL_746067 | ···················· | ···················· | ···················· | ···················· | ···················· | ···················· |
| Z | 5345 | hCoV-19/USA/UT-UPHL-2012944794/2020\|EPI_ISL_746068 | ···················· | ···················· | ···················· | ···················· | ···················· | ···················· |
| Z | 5346 | hCoV-19/USA/UT-UPHL-2012787592/2020\|EPI_ISL_746069 | ···················· | ·A·················· | ···················· | ···················· | ···················· | ···················· |
| Z | 5347 | hCoV-19/USA/UT-UPHL-2012080934/2020\|EPI_ISL_746070 | ···················· | ···················· | ···················· | ···················· | ···················· | ···················· |
| Z | 5348 | hCoV-19/USA/UT-UPHL-2012933745/2020\|EPI_ISL_746071 | ···················· | ···················· | ···················· | ···················· | ···················· | ···················· |
| Z | 5349 | hCoV-19/USA/UT-UPHL-2012956355/2020\|EPI_ISL_746072 | ···················· | ···················· | ···················· | ···················· | ···················· | ···················· |
| Z | 5350 | hCoV-19/USA/UT-UPHL-2012220745/2020\|EPI_ISL_746073 | ···················· | ···················· | ···················· | ···················· | ···················· | ···················· |
| Z | 5351 | hCoV-19/USA/UT-UPHL-2012411390/2020\|EPI_ISL_746074 | ···················· | ···················· | ···················· | ···················· | ···················· | ···················· |
| Z | 5352 | hCoV-19/USA/UT-UPHL-2012807681/2020\|EPI_ISL_746075 | ···················· | ···················· | ···················· | ···················· | ···················· | ···················· |
| Z | 5353 | hCoV-19/USA/UT-UPHL-2012897929/2020\|EPI_ISL_746076 | ···················· | ···················· | ···················· | ···················· | ···················· | ···················· |
| Z | 5354 | hCoV-19/USA/UT-UPHL-2012988511/2020\|EPI_ISL_746077 | ···················· | ·A·················· | ···················· | ···················· | ···················· | ···················· |
| Z | 5355 | hCoV-19/USA/UT-UPHL-2012903953/2020\|EPI_ISL_746078 | ···················· | ···················· | ···················· | ···················· | ···················· | ···················· |
| Z | 5356 | hCoV-19/USA/UT-UPHL-2012372658/2020\|EPI_ISL_746079 | ···················· | ···················· | ···················· | ···················· | ···················· | ···················· |
| Z | 5357 | hCoV-19/USA/UT-UPHL-2012513092/2020\|EPI_ISL_746080 | ···················· | ···················· | ···················· | ···················· | ···················· | ···················· |
| Z | 5358 | hCoV-19/USA/UT-UPHL-2012382383/2020\|EPI_ISL_746081 | ···················· | ···················· | ···················· | ···················· | ···················· | ···················· |
| Z | 5359 | hCoV-19/USA/UT-UPHL-2012051456/2020\|EPI_ISL_746082 | ···················· | ···················· | ···················· | ···················· | ···················· | ···················· |
| Z | 5360 | hCoV-19/USA/UT-UPHL-2012970003/2020\|EPI_ISL_746083 | ···················· | ···················· | ···················· | ···················· | ···················· | ···················· |
| Z | 5361 | hCoV-19/USA/UT-UPHL-2012298741/2020\|EPI_ISL_746084 | ···················· | ···················· | ···················· | ···················· | ···················· | ···················· |
| Z | 5362 | hCoV-19/USA/UT-UPHL-2012304282/2020\|EPI_ISL_746085 | ···················· | ···················· | ···················· | ···················· | ···················· | ···················· |
| Z | 5363 | hCoV-19/USA/UT-UPHL-2012495146/2020\|EPI_ISL_746086 | ···················· | ···················· | ···················· | ···················· | ···················· | ···················· |
| Z | 5364 | hCoV-19/USA/UT-UPHL-2012972277/2020\|EPI_ISL_746087 | ···················· | ·A·················· | ···················· | ···················· | ···················· | ···················· |
| Z | 5365 | hCoV-19/USA/UT-UPHL-2012579569/2020\|EPI_ISL_746088 | ···················· | ·A·················· | ···················· | ···················· | ···················· | ···················· |
| Z | 5366 | hCoV-19/USA/UT-UPHL-2012490016/2020\|EPI_ISL_746089 | ···················· | ·A·················· | ···················· | ···················· | ···················· | ···················· |
| Z | 5367 | hCoV-19/USA/UT-UPHL-2012776941/2020\|EPI_ISL_746090 | ···················· | ···················· | ···················· | ···················· | ···················· | ···················· |
| Z | 5368 | hCoV-19/USA/UT-UPHL-2012343351/2020\|EPI_ISL_746091 | ···················· | ···················· | ···················· | ···················· | ···················· | ···················· |
| Z | 5369 | hCoV-19/USA/UT-UPHL-2012734844/2020\|EPI_ISL_746092 | ···················· | ···················· | ···················· | ···················· | ···················· | ···················· |
| Z | 5370 | hCoV-19/USA/UT-UPHL-2012159552/2020\|EPI_ISL_746093 | ···················· | ···················· | ···················· | ···················· | ···················· | ···················· |
| Z | 5371 | hCoV-19/USA/UT-UPHL-2012026968/2020\|EPI_ISL_746094 | ···················· | ·A·················· | ···················· | ···················· | ···················· | ···················· |
| Z | 5372 | hCoV-19/USA/UT-UPHL-2012753967/2020\|EPI_ISL_746095 | ···················· | ···················· | ···················· | ···················· | ···················· | ···················· |
| Z | 5373 | hCoV-19/USA/UT-UPHL-2012112412/2020\|EPI_ISL_746096 | ···················· | ·A····A············· | ···················· | ···················· | ···················· | ···················· |
| Z | 5374 | hCoV-19/USA/UT-UPHL-2012609524/2020\|EPI_ISL_746097 | ···················· | ·A·················· | ···················· | ···················· | ···················· | ···················· |
| Z | 5375 | hCoV-19/USA/UT-UPHL-2012266151/2020\|EPI_ISL_746098 | ···················· | ···················· | ···················· | ···················· | ···················· | ···················· |
| Z | 5376 | hCoV-19/USA/UT-UPHL-2012828939/2020\|EPI_ISL_746099 | ···················· | ···················· | ···················· | ···················· | ···················· | ···················· |
| Z | 5377 | hCoV-19/USA/CA-CZB-15325/2020\|EPI_ISL_739020\|2020- | ···················· | ·A·················· | ···················· | ···················· | ···················· | ···················· |
| Z | 5378 | hCoV-19/USA/CA-CZB-14107/2020\|EPI_ISL_739029\|2020- | ···················· | ···················· | ···················· | ···················· | ···················· | ···················· |
| Z | 5379 | hCoV-19/USA/CA-CZB-15223/2020\|EPI_ISL_739030\|2020- | ···················· | ···················· | ···················· | ···················· | ···················· | ···················· |
| Z | 5381 | hCoV-19/USA/CA-CZB-14792/2020\|EPI_ISL_739032\|2020- | ···················· | ···················· | ···················· | ···················· | ···················· | ···················· |
| Z | 5382 | hCoV-19/USA/CA-CZB-13766/2020\|EPI_ISL_739033\|2020- | ···················· | ·A·················· | ···················· | ···················· | ···················· | ···················· |
| Z | 5383 | hCoV-19/USA/CA-CZB-14935/2020\|EPI_ISL_739034\|2020- | ···················· | ···················· | ···················· | ···················· | ···················· | ···················· |
| Z | 5385 | hCoV-19/USA/CA-CZB-15984/2020\|EPI_ISL_739036\|2020- | ···················· | ···················· | ···················· | ···················· | ···················· | ···················· |
| Z | 5386 | hCoV-19/USA/CA-CZB-15964/2020\|EPI_ISL_739037\|2020- | ···················· | ···················· | ···················· | ···················· | NN·················· | ···················· |
| Z | 5387 | hCoV-19/USA/CA-CZB-15493/2020\|EPI_ISL_739038\|2020- | ···················· | ·A·················· | ···················· | ···················· | ···················· | ···················· |
| Z | 5388 | hCoV-19/USA/CA-CZB-13698/2020\|EPI_ISL_739039\|2020- | ···················· | ···················· | ···················· | ···················· | ···················· | ···················· |
| Z | 5389 | hCoV-19/Luxembourg/LNS1880857/2020\|EPI_ISL_740010\| | ···················· | ···················· | ···················· | ···················· | ···················· | ···················· |
| Z | 5390 | hCoV-19/Luxembourg/LNS1883126/2020\|EPI_ISL_740011\| | ···················· | ···················· | ···················· | ···················· | ···················· | ···················· |
| Z | 5391 | hCoV-19/Luxembourg/LNS1884814/2020\|EPI_ISL_740012\| | ···················· | ···················· | ···················· | ···················· | ···················· | ···················· |
| Z | 5392 | hCoV-19/Luxembourg/LNS1895902/2020\|EPI_ISL_740013\| | ···················· | ···················· | ···················· | ···················· | ···················· | ···················· |
| Z | 5393 | hCoV-19/Luxembourg/LNS1896192/2020\|EPI_ISL_740014\| | ···················· | ···················· | ···················· | ···················· | ···················· | ···················· |
| Z | 5394 | hCoV-19/Luxembourg/LNS1896881/2020\|EPI_ISL_740015\| | ···················· | ···················· | ···················· | ···················· | ···················· | ···················· |
| Z | 5395 | hCoV-19/Luxembourg/LNS1904080/2020\|EPI_ISL_740017\| | ···················· | ···················· | ···················· | ···················· | ···················· | ···················· |
| Z | 5396 | hCoV-19/Luxembourg/LNS1906480/2020\|EPI_ISL_740018\| | ···················· | ···················· | ···················· | ···················· | ···················· | ···················· |
| Z | 5397 | hCoV-19/Luxembourg/LNS1908675/2020\|EPI_ISL_740019\| | ···················· | ···················· | ···················· | ···················· | ···················· | ···················· |
| Z | 5398 | hCoV-19/Luxembourg/LNS1908786/2020\|EPI_ISL_740020\| | ···················· | ···················· | ···················· | ···················· | ···················· | ···················· |
| Z | 5399 | hCoV-19/Luxembourg/LNS1916011/2020\|EPI_ISL_740021\| | ···················· | ···················· | ···················· | ···················· | ······T············· | ···················· |
| Z | 5400 | hCoV-19/Luxembourg/LNS1916767/2020\|EPI_ISL_740022\| | ···················· | ···················· | ···················· | ···················· | ···················· | ···················· |
| Z | 5401 | hCoV-19/Luxembourg/LNS1930777/2020\|EPI_ISL_740024\| | ···················· | ···················· | ···················· | ···················· | ···················· | ···················· |
| Z | 5402 | hCoV-19/Luxembourg/LNS1931761/2020\|EPI_ISL_740025\| | ···················· | ···················· | ···················· | ···················· | ···················· | ···················· |
| Z | 5403 | hCoV-19/Luxembourg/LNS1932776/2020\|EPI_ISL_740026\| | ···················· | ···················· | ···················· | ···················· | ···················· | ···················· |
| Z | 5404 | hCoV-19/England/LOND-12673BE/2020\|EPI_ISL_741281\|2 | ···················· | ···················· | ···················· | ···················· | ···················· | ···················· |
| Z | 5405 | hCoV-19/England/LOND-12673CD/2020\|EPI_ISL_741282\|2 | ···················· | ·A·················· | ···················· | ···················· | ···················· | ···A················ |
| Z | 5406 | hCoV-19/England/LOND-1267415/2020\|EPI_ISL_741286\|2 | ···················· | ···················· | ···················· | ···················· | ···················· | ···················· |
| Z | 5407 | hCoV-19/England/LOND-1267503/2020\|EPI_ISL_741301\|2 | ···················· | ···················· | ···················· | ···················· | ···················· | ···················· |
| Z | 5408 | hCoV-19/England/LOND-126754F/2020\|EPI_ISL_741302\|2 | ···················· | ···················· | ···················· | ···················· | ···················· | ···················· |
| Z | 5409 | hCoV-19/England/LOND-12E8467/2020\|EPI_ISL_741309\|2 | ···················· | ···················· | ···················· | ···················· | ···················· | ···················· |
| Z | 5410 | hCoV-19/England/LOND-12E8485/2020\|EPI_ISL_741311\|2 | ···················· | ···················· | ···················· | ···················· | ···················· | ···················· |
| Z | 5411 | hCoV-19/England/LOND-12E86CB/2020\|EPI_ISL_741345\|2 | ···················· | ···················· | ···················· | ···················· | ···················· | ···················· |
| Z | 5412 | hCoV-19/England/MILK-A194C2/2020\|EPI_ISL_741347\|20 | ···················· | ···················· | ···················· | ···················· | ···················· | ···················· |
| Z | 5413 | hCoV-19/England/MILK-A23B39/2020\|EPI_ISL_741348\|20 | ···················· | ···················· | ···················· | ···················· | ···················· | ···················· |
| Z | 5414 | hCoV-19/England/MILK-A90FD1/2020\|EPI_ISL_741349\|20 | ···················· | ···················· | ···················· | ···················· | ···················· | ···················· |
| Z | 5415 | hCoV-19/England/MILK-A913D4/2020\|EPI_ISL_741350\|20 | ···················· | ···················· | ···················· | ···················· | ···················· | ···················· |
| Z | 5416 | hCoV-19/England/MILK-A9141D/2020\|EPI_ISL_741351\|20 | ···················· | ···················· | ···················· | ···················· | ···················· | ···················· |
| Z | 5417 | hCoV-19/England/MILK-A914D1/2020\|EPI_ISL_741352\|20 | ···················· | ···················· | ···················· | ···················· | ···················· | ···················· |
| Z | 5418 | hCoV-19/England/MILK-A91A48/2020\|EPI_ISL_741353\|20 | ···················· | ···················· | ···················· | ···················· | ···················· | ···················· |
| Z | 5419 | hCoV-19/England/MILK-A91A57/2020\|EPI_ISL_741354\|20 | ···················· | ···················· | ···················· | ···················· | ···················· | ···················· |
| Z | 5420 | hCoV-19/England/MILK-A91C60/2020\|EPI_ISL_741355\|20 | ···················· | ···················· | ···················· | ···················· | ···················· | ···················· |
| Z | 5421 | hCoV-19/England/MILK-ACFF51/2020\|EPI_ISL_741356\|20 | ···················· | ···················· | ···················· | ···················· | ···················· | ···················· |

| | | | | | | | | |
|---|---|---|---|---|---|---|---|---|
| Z | 5424 | hCoV-19/Wales/PHWC-49C7D3/2020\|EPI_ISL_742604\|2020 | .................... | .................... | ..................... | .................... | .................... | .................... |
| Z | 5425 | hCoV-19/Wales/PHWC-49C7E2/2020\|EPI_ISL_742605\|2020 | .................... | .................... | ..................... | .................... | .................... | .................... |
| Z | 5426 | hCoV-19/Wales/PHWC-49C7F1/2020\|EPI_ISL_742606\|2020 | .................... | .................... | ..................... | .................... | .................... | .................... |
| Z | 5427 | hCoV-19/Wales/PHWC-49C80D/2020\|EPI_ISL_742607\|2020 | .................... | .................... | ..................... | .................... | .................... | .................... |
| Z | 5428 | hCoV-19/Wales/PHWC-49C82B/2020\|EPI_ISL_742608\|2020 | .................... | .................... | ..................... | .................... | .................... | .................... |
| Z | 5429 | hCoV-19/Wales/PHWC-49C849/2020\|EPI_ISL_742610\|2020 | .................... | .................... | ..................... | .................... | .................... | .................... |
| Z | 5430 | hCoV-19/Wales/PHWC-49C858/2020\|EPI_ISL_742611\|2020 | .................... | .................... | ..................... | .................... | .................... | .................... |
| Z | 5431 | hCoV-19/Wales/PHWC-49C867/2020\|EPI_ISL_742612\|2020 | .................... | .................... | ..................... | .................... | .................... | .................... |
| Z | 5432 | hCoV-19/Wales/PHWC-49C8A3/2020\|EPI_ISL_742614\|2020 | .................... | .................... | ..................... | .................... | .................... | .................... |
| Z | 5433 | hCoV-19/Wales/PHWC-49C8C1/2020\|EPI_ISL_742616\|2020 | .................... | .................... | ..................... | .................... | .................... | .................... |
| Z | 5434 | hCoV-19/Wales/PHWC-49C8D0/2020\|EPI_ISL_742617\|2020 | .................... | .................... | ..................... | .................... | .................... | .................... |
| Z | 5435 | hCoV-19/Wales/PHWC-49C8FE/2020\|EPI_ISL_742619\|2020 | .................... | .................... | ..................... | .................... | .................... | .................... |
| Z | 5436 | hCoV-19/Wales/PHWC-49C919/2020\|EPI_ISL_742621\|2020 | .................... | .................... | ..................... | .................... | .................... | .................... |
| Z | 5438 | hCoV-19/Wales/PHWC-49C9A0/2020\|EPI_ISL_742625\|2020 | .................... | .................... | ..................... | .................... | .................... | .................... |
| Z | 5439 | hCoV-19/Wales/PHWC-49C9BF/2020\|EPI_ISL_742626\|2020 | .................... | .................... | ..................... | .................... | .................... | .................... |
| Z | 5440 | hCoV-19/Wales/PHWC-49C9DD/2020\|EPI_ISL_742628\|2020 | .................... | .................... | ..................... | .................... | .................... | .................... |
| Z | 5441 | hCoV-19/Wales/PHWC-49C9EC/2020\|EPI_ISL_742629\|2020 | .................... | .................... | ..................... | .................... | .................... | .................... |
| Z | 5443 | hCoV-19/Wales/PHWC-49CA43/2020\|EPI_ISL_742635\|2020 | .................... | .................... | ..................... | .................... | .................... | .................... |
| Z | 5444 | hCoV-19/Wales/PHWC-49CA52/2020\|EPI_ISL_742636\|2020 | .................... | .................... | ..................... | .................... | .................... | .................... |
| Z | 5446 | hCoV-19/Wales/PHWC-49CA70/2020\|EPI_ISL_742638\|2020 | .................... | .................... | ..................... | .................... | .................... | .................... |
| Z | 5447 | hCoV-19/Wales/PHWC-49CA8F/2020\|EPI_ISL_742639\|2020 | .................... | .................... | ..................... | .................... | .................... | .................... |
| Z | 5449 | hCoV-19/Wales/PHWC-49CAAD/2020\|EPI_ISL_742641\|2020 | .................... | .................... | ..................... | .................... | .................... | .................... |
| Z | 5450 | hCoV-19/Wales/PHWC-49CABC/2020\|EPI_ISL_742642\|2020 | .................... | .................... | ..................... | .................... | .................... | .................... |
| Z | 5451 | hCoV-19/Wales/PHWC-49CADA/2020\|EPI_ISL_742643\|2020 | .................... | .................... | ..................... | .................... | .................... | .................... |
| Z | 5452 | hCoV-19/Wales/PHWC-49CAE9/2020\|EPI_ISL_742644\|2020 | .................... | .................... | ..................... | .................... | .................... | .................... |
| Z | 5453 | hCoV-19/Wales/PHWC-49CC98/2020\|EPI_ISL_742670\|2020 | .................... | .................... | ..................... | .................... | .................... | .................... |
| Z | 5454 | hCoV-19/Wales/PHWC-49CCA7/2020\|EPI_ISL_742671\|2020 | .................... | .................... | ..................... | .................... | .................... | .................... |
| Z | 5455 | hCoV-19/Wales/PHWC-49CCB6/2020\|EPI_ISL_742672\|2020 | .................... | .................... | ..................... | .................... | .................... | .................... |
| Z | 5457 | hCoV-19/Wales/PHWC-49CCE3/2020\|EPI_ISL_742675\|2020 | .................... | .................... | ..................... | .................... | .................... | .................... |
| Z | 5460 | hCoV-19/Wales/PHWC-49CD2C/2020\|EPI_ISL_742679\|2020 | .................... | .................... | ..................... | .................... | .................... | .................... |
| Z | 5462 | hCoV-19/Wales/PHWC-49CD4A/2020\|EPI_ISL_742681\|2020 | .................... | .................... | ..................... | .................... | .................... | .................... |
| Z | 5463 | hCoV-19/Wales/PHWC-49CD59/2020\|EPI_ISL_742682\|2020 | .................... | .................... | ..................... | .................... | .................... | .................... |
| Z | 5464 | hCoV-19/Wales/PHWC-49CD68/2020\|EPI_ISL_742683\|2020 | .................... | .................... | ..................... | .................... | .................... | .................... |
| Z | 5466 | hCoV-19/Wales/PHWC-49CD86/2020\|EPI_ISL_742685\|2020 | .................... | .................... | ..................... | .................... | .................... | .................... |
| Z | 5467 | hCoV-19/Wales/PHWC-49CD95/2020\|EPI_ISL_742686\|2020 | .................... | .................... | ..................... | .................... | .................... | .................... |
| Z | 5468 | hCoV-19/Wales/PHWC-49CDA4/2020\|EPI_ISL_742687\|2020 | .................... | .................... | ..................... | .................... | .................... | .................... |
| Z | 5469 | hCoV-19/Wales/PHWC-49CDB3/2020\|EPI_ISL_742688\|2020 | .................... | .................... | ..................... | .................... | .................... | .................... |
| Z | 5470 | hCoV-19/Wales/PHWC-4A250D/2020\|EPI_ISL_743900\|2020 | .................... | .................... | ..................... | .................... | .................... | .................... |
| Z | 5472 | hCoV-19/Wales/PHWC-4A2549/2020\|EPI_ISL_743904\|2020 | .................... | .................... | ..................... | .................... | .................... | .................... |
| Z | 5473 | hCoV-19/Wales/PHWC-4A2558/2020\|EPI_ISL_743905\|2020 | .................... | .................... | ..................... | .................... | .................... | .................... |
| Z | 5474 | hCoV-19/Wales/PHWC-4A2567/2020\|EPI_ISL_743906\|2020 | .................... | .................... | ..................... | .................... | .................... | .................... |
| Z | 5475 | hCoV-19/Wales/PHWC-4A2576/2020\|EPI_ISL_743907\|2020 | .................... | .................... | ..................... | .................... | .................... | .................... |
| Z | 5476 | hCoV-19/Wales/PHWC-4A2585/2020\|EPI_ISL_743908\|2020 | .................... | .................... | ..................... | .................... | .................... | .................... |
| Z | 5477 | hCoV-19/Wales/PHWC-4A2594/2020\|EPI_ISL_743909\|2020 | .................... | .................... | ..................... | .................... | .................... | .................... |
| Z | 5478 | hCoV-19/Wales/PHWC-4A25A3/2020\|EPI_ISL_743910\|2020 | .................... | .................... | ..................... | .................... | .................... | .................... |
| Z | 5479 | hCoV-19/Wales/PHWC-4A25D0/2020\|EPI_ISL_743913\|2020 | .................... | .................... | ..................... | .................... | .................... | .................... |
| Z | 5480 | hCoV-19/Wales/PHWC-4A25EF/2020\|EPI_ISL_743914\|2020 | .................... | .................... | ..................... | .................... | .................... | .................... |
| Z | 5481 | hCoV-19/Wales/PHWC-4A25FE/2020\|EPI_ISL_743915\|2020 | .................... | .................... | ..................... | .................... | .................... | .................... |
| Z | 5482 | hCoV-19/Wales/PHWC-4A260A/2020\|EPI_ISL_743916\|2020 | .................... | .................... | ..................... | .................... | .................... | .................... |
| Z | 5483 | hCoV-19/Wales/PHWC-4A2619/2020\|EPI_ISL_743917\|2020 | .................... | .................... | ..................... | .................... | .................... | .................... |
| Z | 5484 | hCoV-19/Wales/PHWC-4A2628/2020\|EPI_ISL_743918\|2020 | .................... | .................... | ..................... | .................... | .................... | .................... |
| Z | 5485 | hCoV-19/Wales/PHWC-4A2637/2020\|EPI_ISL_743919\|2020 | .................... | .................... | ..................... | .................... | .................... | .................... |
| Z | 5486 | hCoV-19/Wales/PHWC-4A2646/2020\|EPI_ISL_743920\|2020 | .................... | .................... | ..................... | .................... | .................... | .................... |
| Z | 5487 | hCoV-19/Wales/PHWC-4A2655/2020\|EPI_ISL_743921\|2020 | .................... | .................... | ..................... | .................... | .................... | .................... |
| Z | 5488 | hCoV-19/Wales/PHWC-4A2664/2020\|EPI_ISL_743922\|2020 | .................... | .................... | ..................... | .................... | .................... | .................... |
| Z | 5489 | hCoV-19/Wales/PHWC-4A2682/2020\|EPI_ISL_743924\|2020 | .................... | .................... | ..................... | .................... | .................... | .................... |
| Z | 5490 | hCoV-19/Wales/PHWC-4A2691/2020\|EPI_ISL_743925\|2020 | .................... | .................... | ..................... | .................... | .................... | .................... |
| Z | 5491 | hCoV-19/Wales/PHWC-4A26A0/2020\|EPI_ISL_743926\|2020 | .................... | .................... | ..................... | .................... | .................... | .................... |
| Z | 5492 | hCoV-19/Wales/PHWC-4A26BF/2020\|EPI_ISL_743927\|2020 | .................... | .................... | ..................... | .................... | .................... | .................... |
| Z | 5493 | hCoV-19/Wales/PHWC-4A26CE/2020\|EPI_ISL_743928\|2020 | .................... | .................... | ..................... | .................... | .................... | .................... |
| Z | 5494 | hCoV-19/Wales/PHWC-4A26DD/2020\|EPI_ISL_743929\|2020 | .................... | .................... | ..................... | .................... | .................... | .................... |
| Z | 5495 | hCoV-19/Wales/PHWC-4A26EC/2020\|EPI_ISL_743930\|2020 | .................... | .................... | ..................... | .................... | .................... | .................... |
| Z | 5496 | hCoV-19/Wales/PHWC-4A26FB/2020\|EPI_ISL_743931\|2020 | .................... | .................... | ..................... | .................... | .................... | .................... |
| Z | 5497 | hCoV-19/Wales/PHWC-4A2716/2020\|EPI_ISL_743933\|2020 | .................... | .................... | ..................... | .................... | .................... | .................... |
| Z | 5498 | hCoV-19/Wales/PHWC-4A2734/2020\|EPI_ISL_743934\|2020 | .................... | .................... | ..................... | .................... | .................... | .................... |
| Z | 5499 | hCoV-19/Wales/PHWC-4A2752/2020\|EPI_ISL_743936\|2020 | .................... | .................... | ..................... | .................... | .................... | .................... |
| Z | 5500 | hCoV-19/Wales/PHWC-4A278F/2020\|EPI_ISL_743937\|2020 | .................... | .................... | ..................... | .................... | .................... | .................... |
| Z | 5501 | hCoV-19/Wales/PHWC-4A279E/2020\|EPI_ISL_743938\|2020 | .................... | .................... | ..................... | .................... | .................... | .................... |
| Z | 5502 | hCoV-19/Wales/PHWC-4A27AD/2020\|EPI_ISL_743939\|2020 | .................... | .................... | ..................... | .................... | .................... | .................... |
| Z | 5503 | hCoV-19/Wales/PHWC-4A27BC/2020\|EPI_ISL_743940\|2020 | .................... | .................... | ..................... | .................... | .................... | .................... |
| Z | 5504 | hCoV-19/Wales/PHWC-4A27CB/2020\|EPI_ISL_743941\|2020 | .................... | .................... | ..................... | .................... | .................... | .................... |
| Z | 5505 | hCoV-19/Wales/PHWC-4A27DA/2020\|EPI_ISL_743942\|2020 | .................... | .................... | ..................... | .................... | .................... | .................... |
| Z | 5506 | hCoV-19/Wales/PHWC-4A27E9/2020\|EPI_ISL_743943\|2020 | .................... | .................... | ..................... | .................... | .................... | .................... |
| Z | 5507 | hCoV-19/Wales/PHWC-4A27F8/2020\|EPI_ISL_743944\|2020 | .................... | .................... | ..................... | .................... | .................... | .................... |
| Z | 5508 | hCoV-19/Wales/PHWC-4A2822/2020\|EPI_ISL_743946\|2020 | .................... | .................... | ..................... | .................... | .................... | .................... |
| Z | 5509 | hCoV-19/Wales/PHWC-4A2831/2020\|EPI_ISL_743947\|2020 | .................... | .................... | ..................... | .................... | .................... | .................... |
| Z | 5510 | hCoV-19/Wales/PHWC-4A2840/2020\|EPI_ISL_743948\|2020 | .................... | .................... | ..................... | .................... | .................... | .................... |
| Z | 5511 | hCoV-19/Wales/PHWC-4A285F/2020\|EPI_ISL_743949\|2020 | .................... | .................... | ..................... | .................... | .................... | .................... |
| Z | 5512 | hCoV-19/Wales/PHWC-4A286E/2020\|EPI_ISL_743950\|2020 | .................... | .................... | ..................... | .................... | .................... | .................... |
| Z | 5513 | hCoV-19/Wales/PHWC-4A289B/2020\|EPI_ISL_743952\|2020 | .................... | .................... | ..................... | .................... | .................... | .................... |

| | | | | | | | |
|---|---|---|---|---|---|---|---|
| Z | 5514 hCoV-19/Wales/PHWC-4A28AA/2020\|EPI_ISL_743953\|2020 | ···················· | ···················· | ···················· | ···················· | ···················· | ···················· |
| Z | 5515 hCoV-19/Wales/PHWC-4A28B9/2020\|EPI_ISL_743954\|2020 | ···················· | ······A············· | ···················· | ···················· | ···················· | ···················· |
| Z | 5516 hCoV-19/Wales/PHWC-4A2901/2020\|EPI_ISL_743956\|2020 | ···················· | ···················· | ···················· | ···················· | ···················· | ···················· |
| Z | 5517 hCoV-19/Wales/PHWC-4A2910/2020\|EPI_ISL_743957\|2020 | ···················· | ···················· | ···················· | ···················· | ···················· | ···················· |
| Z | 5518 hCoV-19/Wales/PHWC-4A293E/2020\|EPI_ISL_743958\|2020 | ···················· | ···················· | ···················· | ···················· | ··N················· | ···················· |
| Z | 5519 hCoV-19/Wales/PHWC-4A294D/2020\|EPI_ISL_743959\|2020 | ···················· | ···················· | ···················· | ···················· | ···················· | ···················· |
| Z | 5520 hCoV-19/Wales/PHWC-4A295C/2020\|EPI_ISL_743960\|2020 | ···················· | ···················· | ···················· | ···················· | ···················· | ···················· |
| Z | 5521 hCoV-19/Wales/PHWC-4A2989/2020\|EPI_ISL_743961\|2020 | ···················· | ···················· | ···················· | ···················· | ···················· | ···················· |
| Z | 5522 hCoV-19/Wales/PHWC-4A2998/2020\|EPI_ISL_743962\|2020 | ···················· | ···················· | ···················· | ···················· | ···················· | ···················· |
| Z | 5524 hCoV-19/Wales/PHWC-4A2A2C/2020\|EPI_ISL_743968\|2020 | ···················· | ···················· | ···················· | ···················· | ···················· | ···················· |
| Z | 5525 hCoV-19/Wales/PHWC-4A2A3B/2020\|EPI_ISL_743969\|2020 | ···················· | ···················· | ···················· | ···················· | ···················· | ···················· |
| Z | 5526 hCoV-19/Wales/PHWC-4A2A4A/2020\|EPI_ISL_743970\|2020 | ···················· | ···················· | ···················· | ···················· | ···················· | ···················· |
| Z | 5527 hCoV-19/Wales/PHWC-4A2A59/2020\|EPI_ISL_743971\|2020 | ···················· | ···················· | ···················· | ···················· | ···················· | ···················· |
| Z | 5528 hCoV-19/Wales/PHWC-4A2A68/2020\|EPI_ISL_743972\|2020 | ···················· | ···················· | ···················· | ···················· | ···················· | ···················· |
| Z | 5529 hCoV-19/Wales/PHWC-4A2A86/2020\|EPI_ISL_743973\|2020 | ···················· | ···················· | ···················· | ···················· | ···················· | ···················· |
| Z | 5530 hCoV-19/Wales/PHWC-4A2AA4/2020\|EPI_ISL_743974\|2020 | ···················· | ···················· | ···················· | ···················· | ···················· | ···················· |
| Z | 5531 hCoV-19/Wales/PHWC-4A2AB3/2020\|EPI_ISL_743975\|2020 | ···················· | ···················· | ···················· | ···················· | ···················· | ···················· |
| Z | 5533 hCoV-19/USA/UT-UPHL-2012412077/2020\|EPI_ISL_746100 | ···················· | ···················· | ···················· | ···················· | ···················· | ···················· |
| Z | 5534 hCoV-19/USA/UT-UPHL-2012532895/2020\|EPI_ISL_746101 | ···················· | ·A·················· | ···················· | ···················· | ···················· | ···················· |
| Z | 5535 hCoV-19/USA/UT-UPHL-2012266977/2020\|EPI_ISL_746102 | ···················· | ···················· | ···················· | ···················· | ···················· | ···················· |
| Z | 5536 hCoV-19/USA/UT-UPHL-2012384502/2020\|EPI_ISL_746103 | ···················· | ·A·················· | ···················· | ···················· | ···················· | ···················· |
| Z | 5537 hCoV-19/USA/UT-UPHL-2012425887/2020\|EPI_ISL_746104 | ···················· | ···················· | ···················· | ············A······· | ···················· | ···················· |
| Z | 5538 hCoV-19/USA/UT-UPHL-2012550958/2020\|EPI_ISL_746105 | ···················· | ·A·················· | ···················· | ···················· | ···················· | ···················· |
| Z | 5539 hCoV-19/USA/UT-UPHL-2012011322/2020\|EPI_ISL_746106 | ···················· | ···················· | ···················· | ···················· | ···················· | ···················· |
| Z | 5540 hCoV-19/USA/UT-UPHL-2012516447/2020\|EPI_ISL_746107 | ···················· | ···················· | ···················· | ···················· | ···················· | ···················· |
| Z | 5541 hCoV-19/USA/UT-UPHL-2012826896/2020\|EPI_ISL_746108 | ···················· | ···················· | ···················· | ···················· | ···················· | ···················· |
| Z | 5542 hCoV-19/USA/UT-UPHL-2012217302/2020\|EPI_ISL_746109 | ···················· | ·A·················· | ···················· | ···················· | ···················· | ···················· |
| Z | 5543 hCoV-19/USA/UT-UPHL-2012542638/2020\|EPI_ISL_746110 | ···················· | ·A···A·············· | ···················· | ···················· | ···················· | ···················· |
| Z | 5544 hCoV-19/USA/UT-UPHL-2012332844/2020\|EPI_ISL_746111 | ···················· | ·A·················· | ···················· | ···················· | ···················· | ···················· |
| Z | 5545 hCoV-19/USA/UT-UPHL-2012947909/2020\|EPI_ISL_746112 | ···················· | ···················· | ···················· | ···················· | ···················· | ···················· |
| Z | 5546 hCoV-19/USA/UT-UPHL-2012119573/2020\|EPI_ISL_746113 | ···················· | ···················· | ···················· | ···················· | ···················· | ···················· |
| Z | 5547 hCoV-19/USA/UT-UPHL-2012663262/2020\|EPI_ISL_746114 | ···················· | ·A·················· | ···················· | ···················· | ···················· | ···················· |
| Z | 5548 hCoV-19/USA/UT-UPHL-2012056251/2020\|EPI_ISL_746115 | ···················· | ···················· | ···················· | ···················· | ···················· | ···················· |
| Z | 5549 hCoV-19/USA/UT-UPHL-2012499188/2020\|EPI_ISL_746116 | ···················· | ···················· | ···················· | ···················· | ···················· | ···················· |
| Z | 5550 hCoV-19/USA/UT-UPHL-2012256876/2020\|EPI_ISL_746117 | ···················· | ···················· | ···················· | ···················· | ···················· | ···················· |
| Z | 5551 hCoV-19/USA/UT-UPHL-2012758551/2020\|EPI_ISL_746118 | ···················· | ·A·················· | ···················· | ···················· | ···················· | ···················· |
| Z | 5552 hCoV-19/USA/UT-UPHL-2012780804/2020\|EPI_ISL_746119 | ···················· | ···················· | ···················· | ···················· | ···················· | ···················· |
| Z | 5553 hCoV-19/USA/UT-UPHL-2012738554/2020\|EPI_ISL_746120 | ···················· | ···················· | ···················· | ···················· | ···················· | ···················· |
| Z | 5554 hCoV-19/USA/UT-UPHL-2012120520/2020\|EPI_ISL_746121 | ···················· | ···················· | ···················· | ···················· | ···················· | ···················· |
| Z | 5555 hCoV-19/USA/UT-UPHL-2012524544/2020\|EPI_ISL_746122 | ···················· | ···················· | ···················· | ···················· | ···················· | ···················· |
| Z | 5556 hCoV-19/USA/UT-UPHL-2012135420/2020\|EPI_ISL_746123 | ···················· | ···················· | ···················· | ···················· | ···················· | ···················· |
| Z | 5557 hCoV-19/USA/UT-UPHL-2012400168/2020\|EPI_ISL_746124 | ···················· | ·A·················· | ···················· | ···················· | ···················· | ···················· |
| Z | 5558 hCoV-19/USA/UT-UPHL-2012802849/2020\|EPI_ISL_746125 | ···················· | ···················· | ···················· | ···················· | ···················· | ···················· |
| Z | 5559 hCoV-19/USA/UT-UPHL-2012955640/2020\|EPI_ISL_746126 | ···················· | ···················· | ···················· | ···················· | ···················· | ···················· |
| Z | 5560 hCoV-19/USA/UT-UPHL-2012459276/2020\|EPI_ISL_746127 | ···················· | ·A···A·············· | ···················· | ···················· | ···················· | ···················· |
| Z | 5561 hCoV-19/USA/UT-UPHL-2012106527/2020\|EPI_ISL_746128 | ···················· | ·A·················· | ···················· | ···················· | ···················· | ···················· |
| Z | 5562 hCoV-19/USA/UT-UPHL-2012095441/2020\|EPI_ISL_746129 | ···················· | ···················· | ···················· | ···················· | ···················· | ···················· |
| Z | 5563 hCoV-19/USA/UT-UPHL-2012535481/2020\|EPI_ISL_746130 | ···················· | ···················· | ···················· | ···················· | ···················· | ···················· |
| Z | 5564 hCoV-19/USA/UT-UPHL-2012854117/2020\|EPI_ISL_746131 | ···················· | ···················· | ···················· | ···················· | ···················· | ···················· |
| Z | 5565 hCoV-19/USA/UT-UPHL-2012433014/2020\|EPI_ISL_746132 | ···················· | ···················· | ···················· | ···················· | ···················· | ···················· |
| Z | 5566 hCoV-19/USA/UT-UPHL-2012013798/2020\|EPI_ISL_746133 | ···················· | ···················· | ···················· | ···················· | ···················· | ···················· |
| Z | 5567 hCoV-19/USA/UT-UPHL-2012147164/2020\|EPI_ISL_746134 | ···················· | ···················· | ···················· | ···················· | ···················· | ···················· |
| Z | 5568 hCoV-19/USA/UT-UPHL-2012533179/2020\|EPI_ISL_746135 | ···················· | ···················· | ···················· | ···················· | ···················· | ···················· |
| Z | 5569 hCoV-19/USA/UT-UPHL-2012263380/2020\|EPI_ISL_746136 | ···················· | ·A·················· | ···················· | ···················· | ···················· | ···················· |
| Z | 5570 hCoV-19/USA/UT-UPHL-2012536667/2020\|EPI_ISL_746137 | ···················· | ···················· | ···················· | ···················· | ···················· | ···················· |
| Z | 5571 hCoV-19/USA/UT-UPHL-2012633389/2020\|EPI_ISL_746138 | ···················· | ·A·················· | ···················· | ···················· | ···················· | ···················· |
| Z | 5572 hCoV-19/USA/UT-UPHL-2012163778/2020\|EPI_ISL_746139 | ···················· | ···················· | ···················· | ···················· | ···················· | ···················· |
| Z | 5573 hCoV-19/USA/UT-UPHL-2012485866/2020\|EPI_ISL_746140 | ···················· | ···················· | ···················· | ···················· | ···················· | ···················· |
| Z | 5574 hCoV-19/USA/UT-UPHL-2012182830/2020\|EPI_ISL_746141 | ···················· | ·A·················· | ···················· | ···················· | ···················· | ···················· |
| Z | 5575 hCoV-19/USA/UT-UPHL-2012909359/2020\|EPI_ISL_746142 | ···················· | ···················· | ···················· | ···················· | ···················· | ···················· |
| Z | 5576 hCoV-19/USA/UT-UPHL-2012450649/2020\|EPI_ISL_746143 | ···················· | ···················· | ···················· | ···················· | ···················· | ···················· |
| Z | 5577 hCoV-19/USA/UT-UPHL-2012094478/2020\|EPI_ISL_746144 | ···················· | ·A·················· | ···················· | ···················· | ···················· | ···················· |
| Z | 5578 hCoV-19/USA/UT-UPHL-2012975721/2020\|EPI_ISL_746145 | ···················· | ···················· | ···················· | ···················· | ···················· | ···················· |
| Z | 5579 hCoV-19/USA/UT-UPHL-2012517618/2020\|EPI_ISL_746146 | ···················· | ···················· | ···················· | ···················· | ···················· | ···················· |
| Z | 5580 hCoV-19/USA/UT-UPHL-2012441898/2020\|EPI_ISL_746147 | ···················· | ···················· | ···················· | ···················· | ···················· | ···················· |
| Z | 5581 hCoV-19/USA/UT-UPHL-2012531355/2020\|EPI_ISL_746148 | ···················· | ···················· | ···················· | ···················· | ···················· | ···················· |
| Z | 5582 hCoV-19/USA/UT-UPHL-2012657489/2020\|EPI_ISL_746149 | ···················· | ·A·················· | ···················· | ···················· | ···················· | ···················· |
| Z | 5583 hCoV-19/USA/UT-UPHL-2012564151/2020\|EPI_ISL_746150 | ···················· | ···················· | ···················· | ···················· | ···················· | ···················· |
| Z | 5584 hCoV-19/USA/UT-UPHL-2012045630/2020\|EPI_ISL_746151 | ···················· | ···················· | ···················· | ···················· | ···················· | ···················· |
| Z | 5585 hCoV-19/USA/UT-UPHL-2012607685/2020\|EPI_ISL_746152 | ···················· | ·A·················· | ···················· | ···················· | ···················· | ···················· |
| Z | 5586 hCoV-19/USA/UT-UPHL-2012468965/2020\|EPI_ISL_746153 | ···················· | ·A·················· | ···················· | ···················· | ···················· | ···················· |
| Z | 5587 hCoV-19/USA/UT-UPHL-2012787431/2020\|EPI_ISL_746154 | ···················· | ·A·················· | ···················· | ···················· | ···················· | ···················· |
| Z | 5588 hCoV-19/USA/UT-UPHL-2012040579/2020\|EPI_ISL_746155 | ···················· | ···················· | ···················· | ···················· | ···················· | ···················· |
| Z | 5589 hCoV-19/USA/UT-UPHL-2012381065/2020\|EPI_ISL_746156 | ···················· | ···················· | ···················· | ···················· | ···················· | ···················· |
| Z | 5590 hCoV-19/USA/UT-UPHL-2012547628/2020\|EPI_ISL_746157 | ···················· | ···················· | ···················· | ···················· | ···················· | ···················· |
| Z | 5591 hCoV-19/USA/UT-UPHL-2012214216/2020\|EPI_ISL_746158 | ···················· | ···················· | ···················· | ···················· | ···················· | ···················· |
| Z | 5592 hCoV-19/USA/UT-UPHL-2012209457/2020\|EPI_ISL_746159 | ···················· | ···················· | ···················· | ···················· | ···················· | ···················· |
| Z | 5593 hCoV-19/USA/UT-UPHL-2012589137/2020\|EPI_ISL_746160 | ···················· | ···················· | ···················· | ···················· | ···················· | ···················· |
| Z | 5594 hCoV-19/USA/UT-UPHL-2012787729/2020\|EPI_ISL_746161 | ···················· | ···················· | ···················· | ···················· | ···················· | ···················· |
| Z | 5595 hCoV-19/USA/UT-UPHL-2012734882/2020\|EPI_ISL_746162 | ···················· | ·A·················· | ···················· | ···················· | ···················· | ···················· |

| | | | | | | | | |
|---|---|---|---|---|---|---|---|---|
| Z | 5596 | hCoV-19/USA/UT-UPHL-2012432064/2020\|EPI_ISL_746163 | ···················· | ·A·················· | ···················· | ···················· | ···················· | ···················· |
| Z | 5597 | hCoV-19/USA/UT-UPHL-2012113110/2020\|EPI_ISL_746164 | ···················· | ···················· | ···················· | ···················· | ···················· | ···················· |
| Z | 5598 | hCoV-19/USA/UT-UPHL-2012091811/2020\|EPI_ISL_746165 | ···················· | ···················· | ···················· | ···················· | ···················· | ···················· |
| Z | 5599 | hCoV-19/USA/UT-UPHL-2012908484/2020\|EPI_ISL_746166 | ···················· | ···················· | ···················· | ···················· | ···················· | ···················· |
| Z | 5600 | hCoV-19/USA/UT-UPHL-2012150066/2020\|EPI_ISL_746167 | ···················· | ·A·················· | ···················· | ···················· | ···················· | ···················· |
| Z | 5601 | hCoV-19/USA/UT-UPHL-2012676927/2020\|EPI_ISL_746168 | ···················· | ···················· | ···················· | ···················· | ···················· | ···················· |
| Z | 5602 | hCoV-19/USA/UT-UPHL-2012130118/2020\|EPI_ISL_746169 | ···················· | ·A·················· | ···················· | ···················· | ···················· | ···················· |
| Z | 5603 | hCoV-19/USA/UT-UPHL-2012929325/2020\|EPI_ISL_746170 | ···················· | ···················· | ···················· | ···················· | ···················· | ···················· |
| Z | 5604 | hCoV-19/USA/UT-UPHL-2012551252/2020\|EPI_ISL_746171 | ···················· | ·A·················· | ···················· | ···················· | ···················· | ···················· |
| Z | 5605 | hCoV-19/USA/UT-UPHL-2012067167/2020\|EPI_ISL_746172 | ···················· | ···················· | ···················· | ···················· | ···················· | ···················· |
| Z | 5606 | hCoV-19/USA/UT-UPHL-2012955167/2020\|EPI_ISL_746173 | ···················· | ···················· | ···················· | ···················· | ···················· | ···················· |
| Z | 5607 | hCoV-19/USA/UT-UPHL-2012145654/2020\|EPI_ISL_746174 | ···················· | ···················· | ···················· | ···················· | ···················· | ···················· |
| Z | 5608 | hCoV-19/USA/UT-UPHL-2012013391/2020\|EPI_ISL_746175 | ···················· | ···················· | ···················· | ···················· | ···················· | ···················· |
| Z | 5609 | hCoV-19/USA/UT-UPHL-2012977576/2020\|EPI_ISL_746176 | ···················· | ···················· | ···················· | ···················· | ···················· | ···················· |
| Z | 5610 | hCoV-19/USA/UT-UPHL-2012513757/2020\|EPI_ISL_746177 | ···················· | ···················· | ···················· | ···················· | ···················· | ···················· |
| Z | 5611 | hCoV-19/USA/UT-UPHL-2012168362/2020\|EPI_ISL_746178 | ···················· | ···················· | ···················· | ···················· | ···················· | ···················· |
| Z | 5612 | hCoV-19/USA/UT-UPHL-2012086479/2020\|EPI_ISL_746179 | ···················· | ···················· | ···················· | ···················· | ···················· | ···················· |
| Z | 5613 | hCoV-19/USA/UT-UPHL-2012214394/2020\|EPI_ISL_746180 | ···················· | ···················· | ···················· | ···················· | ···················· | ···················· |
| Z | 5614 | hCoV-19/USA/UT-UPHL-2012248379/2020\|EPI_ISL_746181 | ···················· | ···················· | ···················· | ···················· | ···················· | ···················· |
| Z | 5615 | hCoV-19/USA/UT-UPHL-2012106502/2020\|EPI_ISL_746182 | ···················· | ···················· | ···················· | ···················· | ···················· | ···················· |
| Z | 5616 | hCoV-19/USA/UT-UPHL-2012444066/2020\|EPI_ISL_746183 | ···················· | ···················· | ···················· | ···················· | ···················· | ···················· |
| Z | 5617 | hCoV-19/USA/UT-UPHL-2012046073/2020\|EPI_ISL_746184 | ···················· | ···················· | ···················· | ···················· | ···················· | ···················· |
| Z | 5618 | hCoV-19/USA/UT-UPHL-2012496161/2020\|EPI_ISL_746185 | ···················· | ·A·················· | ···················· | ···················· | ···················· | ···················· |
| Z | 5619 | hCoV-19/USA/UT-UPHL-2012035600/2020\|EPI_ISL_746186 | ···················· | ···················· | ···················· | ···················· | ···················· | ···················· |
| Z | 5620 | hCoV-19/USA/UT-UPHL-2012437782/2020\|EPI_ISL_746187 | ···················· | ···················· | ···················· | ···················· | ···················· | ···················· |
| Z | 5621 | hCoV-19/USA/UT-UPHL-2012395205/2020\|EPI_ISL_746188 | ···················· | ···················· | ···················· | ···················· | ···················· | ···················· |
| Z | 5622 | hCoV-19/USA/UT-UPHL-2012361798/2020\|EPI_ISL_746189 | ···················· | ···················· | ···················· | ···················· | ···················· | ···················· |
| Z | 5623 | hCoV-19/USA/UT-UPHL-2012241339/2020\|EPI_ISL_746190 | ···················· | ·A·················· | ···················· | ···················· | ···················· | ···················· |
| Z | 5624 | hCoV-19/USA/UT-UPHL-2012038375/2020\|EPI_ISL_746191 | ···················· | ···················· | ···················· | ···················· | ···················· | ···················· |
| Z | 5625 | hCoV-19/USA/UT-UPHL-2012619534/2020\|EPI_ISL_746192 | ···················· | ···················· | ···················· | ···················· | ···················· | ···················· |
| Z | 5626 | hCoV-19/USA/UT-UPHL-2012729714/2020\|EPI_ISL_746193 | ···················· | ·A·················· | ···················· | ···················· | ···················· | ···················· |
| Z | 5627 | hCoV-19/USA/UT-UPHL-2012649796/2020\|EPI_ISL_746194 | ···················· | ·A·················· | ···················· | ···················· | ···················· | ···················· |
| Z | 5628 | hCoV-19/USA/UT-UPHL-2012872257/2020\|EPI_ISL_746195 | ···················· | ···················· | ···················· | ···················· | ···················· | ···················· |
| Z | 5629 | hCoV-19/USA/UT-UPHL-2012309369/2020\|EPI_ISL_746196 | ···················· | ···················· | ···················· | ···················· | ···················· | ···················· |
| Z | 5630 | hCoV-19/USA/UT-UPHL-2012622027/2020\|EPI_ISL_746197 | ···················· | ···················· | ···················· | ···················· | ···················· | ···················· |
| Z | 5631 | hCoV-19/USA/UT-UPHL-2012432369/2020\|EPI_ISL_746198 | ···················· | ·A·················· | ···················· | ···················· | ···················· | ···················· |
| Z | 5632 | hCoV-19/USA/UT-UPHL-2012580849/2020\|EPI_ISL_746199 | ···················· | ···················· | ···················· | ···················· | ···················· | ···················· |
| Z | 5633 | hCoV-19/USA/CA-CZB-14812/2020\|EPI_ISL_739000\|2020- | ···················· | ···················· | ···················· | ···················· | ···················· | ···················· |
| Z | 5634 | hCoV-19/USA/CA-CZB-14817/2020\|EPI_ISL_739001\|2020- | ···················· | ···················· | ···················· | ···················· | ···················· | ···················· |
| Z | 5635 | hCoV-19/USA/CA-CZB-15434/2020\|EPI_ISL_739002\|2020- | ···················· | ···················· | ···················· | ···················· | ···················· | ···················· |
| Z | 5636 | hCoV-19/USA/CA-CZB-14118/2020\|EPI_ISL_739003\|2020- | ···················· | ···················· | ···················· | ···················· | ···················· | ···················· |
| Z | 5638 | hCoV-19/USA/CA-CZB-15255/2020\|EPI_ISL_739005\|2020- | ···················· | ···················· | ···················· | ···················· | ···················· | ···················· |
| Z | 5639 | hCoV-19/USA/CA-CZB-13685/2020\|EPI_ISL_739006\|2020- | ···················· | ···················· | ···················· | ···················· | ···················· | ···················· |
| Z | 5640 | hCoV-19/USA/CA-CZB-14819/2020\|EPI_ISL_739007\|2020- | ···················· | ···················· | ···················· | ···················· | ···················· | ···················· |
| Z | 5641 | hCoV-19/USA/CA-CZB-15153/2020\|EPI_ISL_739008\|2020- | ···················· | ·A·················· | ···················· | ···················· | ···················· | ···················· |
| Z | 5642 | hCoV-19/USA/CA-CZB-15959/2020\|EPI_ISL_739009\|2020- | ···················· | ···················· | ···················· | ···················· | ···················· | ···················· |
| Z | 5643 | hCoV-19/USA/CA-CZB-14547/2020\|EPI_ISL_739010\|2020- | ···················· | ·A·················· | ···················· | ···················· | ···················· | ···················· |
| Z | 5644 | hCoV-19/USA/CA-CZB-14717/2020\|EPI_ISL_739011\|2020- | ···················· | ···················· | ···················· | ···················· | ·········A·········· | ···················· |
| Z | 5645 | hCoV-19/USA/CA-CZB-15121/2020\|EPI_ISL_739012\|2020- | ···················· | ···················· | ···················· | ···················· | ···················· | ···················· |
| Z | 5646 | hCoV-19/USA/CA-CZB-14899/2020\|EPI_ISL_739013\|2020- | ···················· | ···················· | ···················· | ···················· | ···················· | ···················· |
| Z | 5647 | hCoV-19/USA/CA-CZB-16176/2020\|EPI_ISL_739014\|2020- | ···················· | ···················· | ···················· | ···················· | ···················· | ···················· |
| Z | 5648 | hCoV-19/USA/CA-CZB-14581/2020\|EPI_ISL_739015\|2020- | ···················· | ···················· | ···················· | ···················· | ···················· | ···················· |
| Z | 5649 | hCoV-19/USA/CA-CZB-14227/2020\|EPI_ISL_739016\|2020- | ···················· | ···················· | ···················· | ···················· | ···················· | ···················· |
| Z | 5650 | hCoV-19/USA/CA-CZB-15494/2020\|EPI_ISL_739017\|2020- | ···················· | ·A·················· | ···················· | ···················· | ···················· | ···················· |
| Z | 5651 | hCoV-19/USA/CA-CZB-14072/2020\|EPI_ISL_739018\|2020- | ···················· | ···················· | ···················· | ···················· | ···················· | ···················· |
| Z | 5652 | hCoV-19/USA/CA-CZB-13681/2020\|EPI_ISL_739019\|2020- | ···················· | ·A·················· | ···················· | ···················· | ···················· | ···················· |
| Z | 5653 | hCoV-19/USA/CA-CZB-15133/2020\|EPI_ISL_739021\|2020- | ···················· | ·A·················· | ···················· | ···················· | ···················· | ···················· |
| Z | 5654 | hCoV-19/USA/CA-CZB-16051/2020\|EPI_ISL_739022\|2020- | ···················· | ·A·················· | ···················· | ···················· | ···················· | ···················· |
| Z | 5655 | hCoV-19/USA/CA-CZB-13852/2020\|EPI_ISL_739023\|2020- | ···················· | ···················· | ···················· | ···················· | ···················· | ···················· |
| Z | 5656 | hCoV-19/USA/CA-CZB-15360/2020\|EPI_ISL_739024\|2020- | ···················· | ···················· | ···················· | ···················· | ···················· | ···················· |
| Z | 5659 | hCoV-19/USA/CA-CZB-15938/2020\|EPI_ISL_739027\|2020- | ···················· | ···················· | ···················· | ···················· | ···················· | ···················· |
| Z | 5660 | hCoV-19/USA/CA-CZB-14868/2020\|EPI_ISL_739028\|2020- | ···················· | ···················· | ···················· | ···················· | ···················· | ···················· |
| Z | 5661 | hCoV-19/USA/CA-CZB-15976/2020\|EPI_ISL_739040\|2020- | ···················· | ·A·················· | ···················· | ···················· | ···················· | ···················· |
| Z | 5663 | hCoV-19/USA/CA-CZB-13726/2020\|EPI_ISL_739042\|2020- | ···················· | ···················· | ···················· | ···················· | ···················· | ···················· |
| Z | 5664 | hCoV-19/USA/CA-CZB-14801/2020\|EPI_ISL_739043\|2020- | ···················· | ···················· | ···················· | ···················· | ···················· | ···················· |
| Z | 5665 | hCoV-19/USA/CA-CZB-14929/2020\|EPI_ISL_739044\|2020- | ···················· | ···················· | ···················· | ···················· | ···················· | ···················· |
| Z | 5666 | hCoV-19/USA/CA-CZB-15277/2020\|EPI_ISL_739045\|2020- | ···················· | ···················· | ···················· | ···················· | ···················· | ···················· |
| Z | 5667 | hCoV-19/USA/CA-CZB-13861/2020\|EPI_ISL_739046\|2020- | ···················· | ···················· | ···················· | ···················· | ···················· | ···················· |
| Z | 5668 | hCoV-19/USA/CA-CZB-14029/2020\|EPI_ISL_739047\|2020- | ···················· | ···················· | ···················· | ···················· | ···················· | ···················· |
| Z | 5669 | hCoV-19/USA/CA-CZB-14686/2020\|EPI_ISL_739048\|2020- | ···················· | ···················· | ···················· | ···················· | ···················· | ···················· |
| Z | 5670 | hCoV-19/USA/CA-CZB-13926/2020\|EPI_ISL_739049\|2020- | ···················· | ···················· | ···················· | ···················· | ···················· | ···················· |
| Z | 5671 | hCoV-19/USA/CA-CZB-15941/2020\|EPI_ISL_739050\|2020- | ···················· | ···················· | ···················· | ···················· | ···················· | ···················· |
| Z | 5672 | hCoV-19/USA/CA-CZB-14767/2020\|EPI_ISL_739051\|2020- | ···················· | ···················· | ···················· | ···················· | ···················· | ···················· |
| Z | 5673 | hCoV-19/USA/CA-CZB-14760/2020\|EPI_ISL_739052\|2020- | ···················· | ···················· | ···················· | ···················· | ···················· | ···················· |
| Z | 5674 | hCoV-19/USA/CA-CZB-15947/2020\|EPI_ISL_739053\|2020- | ···················· | ···················· | ···················· | ···················· | ···················· | ···················· |
| Z | 5675 | hCoV-19/USA/CA-CZB-14064/2020\|EPI_ISL_739054\|2020- | ···················· | ···················· | ···················· | ···················· | ···················· | ···················· |
| Z | 5676 | hCoV-19/USA/CA-CZB-14652/2020\|EPI_ISL_739055\|2020- | ···················· | ···················· | ···················· | ···················· | ···················· | ···················· |
| Z | 5677 | hCoV-19/USA/CA-CZB-13943/2020\|EPI_ISL_739056\|2020- | ···················· | ···················· | ···················· | ···················· | ···················· | ···················· |
| Z | 5679 | hCoV-19/USA/CA-CZB-15939/2020\|EPI_ISL_739058\|2020- | ···················· | ···················· | ···················· | ···················· | ···················· | ···················· |
| Z | 5680 | hCoV-19/USA/CA-CZB-14285/2020\|EPI_ISL_739059\|2020- | ···················· | ·A·················· | ···················· | ···················· | ···················· | ···················· |

| | | | | | | | | |
|---|---|---|---|---|---|---|---|---|
| Z | 5681 | hCoV-19/USA/CA-CZB-15027/2020\|EPI_ISL_739060\|2020- | ···················· | ···················· | ···················· | ···················· | ···················· | ···················· |
| Z | 5682 | hCoV-19/USA/CA-CZB-14156/2020\|EPI_ISL_739061\|2020- | ···················· | ·A·················· | ···················· | ···················· | ···················· | ···················· |
| Z | 5684 | hCoV-19/USA/CA-CZB-14699/2020\|EPI_ISL_739063\|2020- | ···················· | ···················· | ···················· | ···················· | ···················· | ···················· |
| Z | 5687 | hCoV-19/USA/CA-CZB-15218/2020\|EPI_ISL_739066\|2020- | ···················· | ···················· | ···················· | ···················· | ···················· | ···················· |
| Z | 5689 | hCoV-19/USA/CA-CZB-15970/2020\|EPI_ISL_739068\|2020- | ···················· | ·A·················· | ···················· | ···················· | ···················· | ···················· |
| Z | 5690 | hCoV-19/USA/CA-CZB-14934/2020\|EPI_ISL_739069\|2020- | ···················· | ···················· | ···················· | ···················· | ···················· | ···················· |
| Z | 5691 | hCoV-19/USA/CA-CZB-15127/2020\|EPI_ISL_739070\|2020- | ···················· | ···················· | ···················· | ···················· | ···················· | ···················· |
| Z | 5692 | hCoV-19/USA/CA-CZB-14659/2020\|EPI_ISL_739071\|2020- | ···················· | ···················· | ···················· | ···················· | ···················· | ···················· |
| Z | 5693 | hCoV-19/USA/CA-CZB-14877/2020\|EPI_ISL_739072\|2020- | ···················· | ···················· | ···················· | ···················· | ···················· | ···················· |
| Z | 5694 | hCoV-19/USA/CA-CZB-13838/2020\|EPI_ISL_739073\|2020- | ···················· | ···················· | ···················· | ···················· | ···················· | ···················· |
| Z | 5695 | hCoV-19/USA/CA-CZB-14970/2020\|EPI_ISL_739074\|2020- | ···················· | ···················· | ···················· | ···················· | ···················· | ···················· |
| Z | 5696 | hCoV-19/USA/CA-CZB-13970/2020\|EPI_ISL_739075\|2020- | ···················· | ·A·················· | ···················· | ···················· | ···················· | ···················· |
| Z | 5697 | hCoV-19/USA/CA-CZB-15310/2020\|EPI_ISL_739076\|2020- | ···················· | ·A·················· | ···················· | ···················· | ···················· | ···················· |
| Z | 5698 | hCoV-19/USA/CA-CZB-14363/2020\|EPI_ISL_739077\|2020- | ···················· | ···················· | ···················· | ···················· | ···················· | ···················· |
| Z | 5699 | hCoV-19/USA/CA-CZB-14704/2020\|EPI_ISL_739078\|2020- | ···················· | ···················· | ···················· | ···················· | ···················· | ···················· |
| Z | 5700 | hCoV-19/USA/CA-CZB-14764/2020\|EPI_ISL_739079\|2020- | ···················· | ···················· | ···················· | ···················· | ···················· | ···················· |
| Z | 5701 | hCoV-19/USA/CA-CZB-14529/2020\|EPI_ISL_739080\|2020- | ···················· | ···················· | ···················· | ···················· | ···················· | ···················· |
| Z | 5702 | hCoV-19/USA/CA-CZB-15449/2020\|EPI_ISL_739081\|2020- | ···················· | ···················· | ···················· | ···················· | ···················· | ···················· |
| Z | 5703 | hCoV-19/USA/CA-CZB-14305/2020\|EPI_ISL_739082\|2020- | ···················· | ···················· | ···················· | ···················· | ···················· | ···················· |
| Z | 5704 | hCoV-19/USA/CA-CZB-14355/2020\|EPI_ISL_739083\|2020- | ···················· | ···················· | ···················· | ···················· | ···················· | ···················· |
| Z | 5705 | hCoV-19/USA/CA-CZB-14350/2020\|EPI_ISL_739084\|2020- | ···················· | ···················· | ···················· | ···················· | ···················· | ···················· |
| Z | 5706 | hCoV-19/USA/CA-CZB-15105/2020\|EPI_ISL_739085\|2020- | ···················· | ·A·················· | ···················· | ···················· | ···················· | ···················· |
| Z | 5707 | hCoV-19/USA/CA-CZB-14724/2020\|EPI_ISL_739086\|2020- | ···················· | ···················· | ···················· | ···················· | ···················· | ···················· |
| Z | 5708 | hCoV-19/USA/CA-CZB-14429/2020\|EPI_ISL_739090\|2020- | ···················· | ·A·················· | ···················· | ···················· | ···················· | ···················· |
| Z | 5710 | hCoV-19/USA/CA-CZB-14679/2020\|EPI_ISL_739092\|2020- | ···················· | ···················· | ···················· | ···················· | ···················· | ···················· |
| Z | 5711 | hCoV-19/USA/CA-CZB-13935/2020\|EPI_ISL_739093\|2020- | ···················· | ·A·················· | ···················· | ···················· | ···················· | ···················· |
| Z | 5712 | hCoV-19/USA/CA-CZB-14931/2020\|EPI_ISL_739094\|2020- | ···················· | ·A·················· | ···················· | ···················· | ···················· | ···················· |
| Z | 5713 | hCoV-19/USA/CA-CZB-15483/2020\|EPI_ISL_739095\|2020- | ···················· | ·A·················· | ···················· | ···················· | ···················· | ···················· |
| Z | 5715 | hCoV-19/USA/CA-CZB-14894/2020\|EPI_ISL_739097\|2020- | ···················· | ···················· | ···················· | ···················· | ···················· | ···················· |
| Z | 5716 | hCoV-19/USA/CA-CZB-15201/2020\|EPI_ISL_739098\|2020- | ···················· | ···················· | ···················· | ···················· | ···················· | ···················· |
| Z | 5717 | hCoV-19/USA/CA-CZB-14670/2020\|EPI_ISL_739099\|2020- | ···················· | ···················· | ···················· | ···················· | ···················· | ···················· |
| Z | 5718 | hCoV-19/Luxembourg/LNS1828619/2020\|EPI_ISL_740000\| | ···················· | ···················· | ···················· | ···················· | ···················· | ···················· |
| Z | 5719 | hCoV-19/Luxembourg/LNS1829518/2020\|EPI_ISL_740001\| | ···················· | ···················· | ···················· | ···················· | ···················· | ···················· |
| Z | 5720 | hCoV-19/Luxembourg/LNS1834890/2020\|EPI_ISL_740002\| | ···················· | ···················· | ···················· | ···················· | ···················· | ···················· |
| Z | 5721 | hCoV-19/Luxembourg/LNS1840548/2020\|EPI_ISL_740003\| | ············T······· | ···················· | ···················· | ···················· | ···················· | ···················· |
| Z | 5722 | hCoV-19/Luxembourg/LNS1841754/2020\|EPI_ISL_740004\| | ···················· | ···················· | ···················· | ···················· | ···················· | ···················· |
| Z | 5723 | hCoV-19/Luxembourg/LNS1846494/2020\|EPI_ISL_740005\| | ···················· | ···················· | ···················· | ···················· | ···················· | ···················· |
| Z | 5724 | hCoV-19/Luxembourg/LNS1848261/2020\|EPI_ISL_740006\| | ···················· | ···················· | ···················· | ···················· | ···················· | ···················· |
| Z | 5725 | hCoV-19/Luxembourg/LNS1850602/2020\|EPI_ISL_740007\| | ···················· | ···················· | ···················· | ···················· | ···················· | ···················· |
| Z | 5726 | hCoV-19/Luxembourg/LNS1856024/2020\|EPI_ISL_740008\| | ···················· | ···················· | ···················· | ···················· | ···················· | ···················· |
| Z | 5727 | hCoV-19/Luxembourg/LNS1869765/2020\|EPI_ISL_740009\| | ···················· | ···················· | ···················· | ···················· | ···················· | ···················· |
| Z | 5728 | hCoV-19/Luxembourg/LNS1947589/2020\|EPI_ISL_740027\| | ···················· | ···················· | ···················· | ···················· | ···················· | ···················· |
| Z | 5729 | hCoV-19/Luxembourg/LNS1956201/2020\|EPI_ISL_740028\| | ···················· | ···················· | ···················· | ···················· | ···················· | ···················· |
| Z | 5730 | hCoV-19/Luxembourg/LNS1960664/2020\|EPI_ISL_740029\| | ···················· | ···················· | ···················· | ···················· | ···················· | ···················· |
| Z | 5731 | hCoV-19/Luxembourg/LNS1963903/2020\|EPI_ISL_740030\| | ···················· | ···················· | ···················· | ···················· | ···················· | ···················· |
| Z | 5732 | hCoV-19/Luxembourg/LNS1973766/2020\|EPI_ISL_740031\| | ···················· | ···················· | ···················· | ···················· | ···················· | ···················· |
| Z | 5733 | hCoV-19/Luxembourg/LNS1987832/2020\|EPI_ISL_740032\| | ···················· | ···················· | ···················· | ···················· | ···················· | ···················· |
| Z | 5734 | hCoV-19/Luxembourg/LNS2001612/2020\|EPI_ISL_740034\| | ···················· | ···················· | ···················· | ···················· | ···················· | ···················· |
| Z | 5735 | hCoV-19/Luxembourg/LNS2002848/2020\|EPI_ISL_740035\| | ···················· | ···················· | ···················· | ···················· | ···················· | ···················· |
| Z | 5736 | hCoV-19/Luxembourg/LNS2003228/2020\|EPI_ISL_740036\| | ···················· | ···················· | ···················· | ···················· | ···················· | ···················· |
| Z | 5737 | hCoV-19/Luxembourg/LNS2011060/2020\|EPI_ISL_740037\| | ···················· | ···················· | ···················· | ···················· | ···················· | ···················· |
| Z | 5738 | hCoV-19/Luxembourg/LNS2021453/2020\|EPI_ISL_740039\| | ···················· | ···················· | ···················· | ···················· | ···················· | ···················· |
| Z | 5739 | hCoV-19/Luxembourg/LNS2033603/2020\|EPI_ISL_740041\| | ···················· | ···················· | ···················· | ···················· | ···················· | ···················· |
| Z | 5740 | hCoV-19/Luxembourg/LNS2033831/2020\|EPI_ISL_740042\| | ···················· | ···················· | ···················· | ···················· | ···················· | ···················· |
| Z | 5741 | hCoV-19/Luxembourg/LNS2036226/2020\|EPI_ISL_740043\| | ···················· | ···················· | ···················· | ···················· | ···················· | ···················· |
| Z | 5742 | hCoV-19/Luxembourg/LNS2037025/2020\|EPI_ISL_740044\| | ············T······· | ···················· | ···················· | ···················· | ···················· | ···················· |
| Z | 5743 | hCoV-19/Luxembourg/LNS2038651/2020\|EPI_ISL_740045\| | ···················· | ···················· | ···················· | ···················· | ···················· | ···················· |
| Z | 5744 | hCoV-19/Luxembourg/LNS2048098/2020\|EPI_ISL_740046\| | ···················· | ···················· | ···················· | ···················· | ···················· | ···················· |
| Z | 5745 | hCoV-19/Luxembourg/LNS2048899/2020\|EPI_ISL_740047\| | ···················· | ···················· | ···················· | ···················· | ···················· | ···················· |
| Z | 5746 | hCoV-19/Luxembourg/LNS2048918/2020\|EPI_ISL_740048\| | ···················· | ···················· | ···················· | ···················· | ···················· | ···················· |
| Z | 5747 | hCoV-19/Luxembourg/LNS2064422/2020\|EPI_ISL_740051\| | ···················· | ···················· | ···················· | ···················· | ···················· | ···················· |
| Z | 5748 | hCoV-19/Luxembourg/LNS2067807/2020\|EPI_ISL_740052\| | ···················· | ···················· | ···················· | ···················· | ···················· | ···················· |
| Z | 5749 | hCoV-19/Luxembourg/LNS2068754/2020\|EPI_ISL_740053\| | ···················· | ···················· | ···················· | ···················· | ···················· | ···················· |
| Z | 5750 | hCoV-19/Luxembourg/LNS2074154/2020\|EPI_ISL_740054\| | ···················· | ···················· | ···················· | ···················· | ···················· | ···················· |
| Z | 5751 | hCoV-19/Luxembourg/LNS2075302/2020\|EPI_ISL_740055\| | ···················· | ···················· | ···················· | ···················· | ···················· | ···················· |
| Z | 5752 | hCoV-19/Luxembourg/LNS2082727/2020\|EPI_ISL_740056\| | ···················· | ···················· | ···················· | ···················· | ···················· | ···················· |
| Z | 5753 | hCoV-19/Luxembourg/LNS2082730/2020\|EPI_ISL_740057\| | ···················· | ···················· | ···················· | ···················· | ···················· | ···················· |
| Z | 5754 | hCoV-19/Luxembourg/LNS2086754/2020\|EPI_ISL_740058\| | ···················· | ···················· | ···················· | ···················· | ···················· | ···················· |
| Z | 5755 | hCoV-19/Luxembourg/LNS2094950/2020\|EPI_ISL_740059\| | ···················· | ···················· | ···················· | ···················· | ···················· | ···················· |
| Z | 5756 | hCoV-19/Luxembourg/LNS2098672/2020\|EPI_ISL_740060\| | ···················· | ···················· | ···················· | ···················· | ···················· | ···················· |
| Z | 5757 | hCoV-19/Luxembourg/LNS2114865/2020\|EPI_ISL_740061\| | ···················· | ···················· | ···················· | ···················· | ···················· | ···················· |
| Z | 5758 | hCoV-19/Luxembourg/LNS2117299/2020\|EPI_ISL_740062\| | ···················· | ···················· | ···················· | ···················· | ···················· | ···················· |
| Z | 5759 | hCoV-19/Luxembourg/LNS2122915/2020\|EPI_ISL_740063\| | ···················· | ···················· | ···················· | ···················· | ···················· | ···················· |
| Z | 5760 | hCoV-19/Luxembourg/LNS2131206/2020\|EPI_ISL_740064\| | ···················· | ···················· | ···················· | ···················· | ···················· | ···················· |
| Z | 5761 | hCoV-19/Luxembourg/LNS2132885/2020\|EPI_ISL_740065\| | ···················· | ····A··············· | ···················· | ···················· | ···················· | ···················· |
| Z | 5762 | hCoV-19/Luxembourg/LNS2137806/2020\|EPI_ISL_740066\| | ···················· | ···················· | ···················· | ···················· | ···················· | ···················· |
| Z | 5763 | hCoV-19/Luxembourg/LNS2148417/2020\|EPI_ISL_740067\| | ···················· | ···················· | ···················· | ···················· | ···················· | ···················· |
| Z | 5764 | hCoV-19/Luxembourg/LNS2157897/2020\|EPI_ISL_740068\| | ···················· | ···················· | ···················· | ···················· | ···················· | ···················· |
| Z | 5765 | hCoV-19/Luxembourg/LNS2162512/2020\|EPI_ISL_740069\| | ···················· | ···················· | ···················· | ···················· | ···················· | ···················· |
| Z | 5766 | hCoV-19/Luxembourg/LNS2179748/2020\|EPI_ISL_740071\| | ···················· | ···················· | ···················· | ···················· | ···················· | ···················· |

| | | | | | | | | |
|---|---|---|---|---|---|---|---|---|
| Z | 5767 | hCoV-19/Luxembourg/LNS2180340/2020\|EPI_ISL_740072\| | ···················· | ···················· | ···················· | ···················· | ···················· | ···················· |
| Z | 5768 | hCoV-19/Luxembourg/LNS2186059/2020\|EPI_ISL_740073\| | ···················· | ···················· | ···················· | ···················· | ···················· | ···················· |
| Z | 5769 | hCoV-19/Luxembourg/LNS2188212/2020\|EPI_ISL_740074\| | ···················· | ···················· | ···················· | ···················· | ···················· | ···················· |
| Z | 5770 | hCoV-19/Luxembourg/LNS2190207/2020\|EPI_ISL_740075\| | ···················· | G··················· | ···················· | ···················· | ···················· | ···················· |
| Z | 5771 | hCoV-19/Luxembourg/LNS2198752/2020\|EPI_ISL_740076\| | ···················· | ···················· | ···················· | ···················· | ···················· | ···················· |
| Z | 5772 | hCoV-19/Luxembourg/LNS2201077/2020\|EPI_ISL_740077\| | ···················· | ···················· | ···················· | ···················· | ···················· | ···················· |
| Z | 5773 | hCoV-19/Luxembourg/LNS2204274/2020\|EPI_ISL_740078\| | ···················· | ···················· | ···················· | ···················· | ···················· | ···················· |
| Z | 5774 | hCoV-19/Luxembourg/LNS2205106/2020\|EPI_ISL_740079\| | ···················· | ···················· | ···················· | ···················· | ···················· | ···················· |
| Z | 5775 | hCoV-19/Luxembourg/LNS2212205/2020\|EPI_ISL_740080\| | ···················· | ···················· | ···················· | ···················· | ···················· | ···················· |
| Z | 5776 | hCoV-19/Luxembourg/LNS2213321/2020\|EPI_ISL_740081\| | ···················· | ···················· | ···················· | ···················· | ···················· | ···················· |
| Z | 5777 | hCoV-19/Luxembourg/LNS2222126/2020\|EPI_ISL_740082\| | ···················· | ···················· | ···················· | ···················· | ···················· | ···················· |
| Z | 5778 | hCoV-19/Luxembourg/LNS2248814/2020\|EPI_ISL_740083\| | ···················· | G··················· | ···················· | ···················· | ···················· | ···················· |
| Z | 5779 | hCoV-19/Luxembourg/LNS2253004/2020\|EPI_ISL_740084\| | ···················· | ···················· | ···················· | ···················· | ···················· | ···················· |
| Z | 5780 | hCoV-19/Luxembourg/LNS2254226/2020\|EPI_ISL_740085\| | ···················· | ···················· | ···················· | ···················· | ···················· | ···················· |
| Z | 5781 | hCoV-19/Luxembourg/LNS2256437/2020\|EPI_ISL_740086\| | ···················· | ···················· | ···················· | ···················· | ···················· | ···················· |
| Z | 5782 | hCoV-19/Luxembourg/LNS2264925/2020\|EPI_ISL_740087\| | ···················· | ···················· | ···················· | ···················· | ···················· | ···················· |
| Z | 5783 | hCoV-19/Luxembourg/LNS2266073/2020\|EPI_ISL_740088\| | ···················· | ···················· | ···················· | ···················· | ···················· | ···················· |
| Z | 5784 | hCoV-19/Luxembourg/LNS2266551/2020\|EPI_ISL_740089\| | ···················· | ···················· | ···················· | ···················· | ···················· | ···················· |
| Z | 5785 | hCoV-19/Luxembourg/LNS2272431/2020\|EPI_ISL_740090\| | ···················· | ···················· | ···················· | ···················· | ···················· | ···················· |
| Z | 5786 | hCoV-19/Luxembourg/LNS2282331/2020\|EPI_ISL_740091\| | ···················· | ···················· | ···················· | ···················· | ···················· | ···················· |
| Z | 5787 | hCoV-19/Luxembourg/LNS2287197/2020\|EPI_ISL_740093\| | ···················· | ···················· | ···················· | ···················· | ···················· | ···················· |
| Z | 5788 | hCoV-19/Luxembourg/LNS2291163/2020\|EPI_ISL_740094\| | ···················· | ···················· | ···················· | ···················· | ···················· | ···················· |
| Z | 5789 | hCoV-19/Luxembourg/LNS2293608/2020\|EPI_ISL_740095\| | ···················· | ···················· | ···················· | ···················· | ···················· | ···················· |
| Z | 5790 | hCoV-19/Luxembourg/LNS2321488/2020\|EPI_ISL_740096\| | ···················· | ···················· | ···················· | ···················· | ···················· | ···················· |
| Z | 5791 | hCoV-19/Luxembourg/LNS2322897/2020\|EPI_ISL_740097\| | ···················· | ···················· | ···················· | ···················· | ···················· | ···················· |
| Z | 5792 | hCoV-19/Luxembourg/LNS2329912/2020\|EPI_ISL_740099\| | ···················· | ···················· | ···················· | ···················· | ···················· | ···················· |
| Z | 5793 | hCoV-19/England/LOND-12E84B2/2020\|EPI_ISL_741314\|2 | ···················· | ···················· | ···················· | ···················· | ···················· | ···················· |
| Z | 5794 | hCoV-19/England/LOND-12E8528/2020\|EPI_ISL_741321\|2 | ···················· | ···················· | ···················· | ···················· | ···················· | ···················· |
| Z | 5795 | hCoV-19/England/LOND-12E85CE/2020\|EPI_ISL_741331\|2 | ···················· | ···················· | ···················· | ···················· | ···················· | ···················· |
| Z | 5796 | hCoV-19/England/LOND-12E85DD/2020\|EPI_ISL_741332\|2 | ···················· | ···················· | ···················· | ···················· | ···················· | ···················· |
| Z | 5797 | hCoV-19/England/LOND-12E85FB/2020\|EPI_ISL_741334\|2 | ···················· | ···················· | ···················· | ···················· | ···················· | ···················· |
| Z | 5798 | hCoV-19/England/LOND-12E8625/2020\|EPI_ISL_741336\|2 | ···················· | ···················· | ···················· | ···················· | ···················· | ···················· |
| Z | 5799 | hCoV-19/England/MILK-B3912E/2020\|EPI_ISL_741359\|20 | ···················· | ···················· | ···················· | ···················· | ···················· | ···················· |
| Z | 5800 | hCoV-19/England/MILK-B39908/2020\|EPI_ISL_741360\|20 | ···················· | ···················· | ···················· | ···················· | ···················· | ···················· |
| Z | 5801 | hCoV-19/England/MILK-B3BAE5/2020\|EPI_ISL_741362\|20 | ···················· | ···················· | ···················· | ···················· | ···················· | ···················· |
| Z | 5802 | hCoV-19/England/MILK-B3CE15/2020\|EPI_ISL_741363\|20 | ···················· | ···················· | ···················· | ···················· | ···················· | ···················· |
| Z | 5803 | hCoV-19/England/MILK-B3CE60/2020\|EPI_ISL_741364\|20 | ···················· | ···················· | ···················· | ···················· | ···················· | ···················· |
| Z | 5804 | hCoV-19/England/MILK-B3D37F/2020\|EPI_ISL_741365\|20 | ···················· | ···················· | ···················· | ···················· | ···················· | ···················· |
| Z | 5805 | hCoV-19/England/MILK-B3D38E/2020\|EPI_ISL_741366\|20 | ···················· | ···················· | ···················· | ···················· | ···················· | ···················· |
| Z | 5806 | hCoV-19/England/MILK-B3D3BB/2020\|EPI_ISL_741367\|20 | ···················· | ···················· | ···················· | ···················· | ···················· | ···················· |
| Z | 5807 | hCoV-19/England/MILK-B3D3F7/2020\|EPI_ISL_741368\|20 | ···················· | ···················· | ···················· | ···················· | ······T············· | ···················· |
| Z | 5808 | hCoV-19/England/MILK-B3D403/2020\|EPI_ISL_741369\|20 | ···················· | ···················· | ···················· | ···················· | ···················· | ···················· |
| Z | 5809 | hCoV-19/England/MILK-B3D412/2020\|EPI_ISL_741370\|20 | ···················· | ···················· | ···················· | ···················· | ···················· | ···················· |
| Z | 5810 | hCoV-19/England/MILK-B3D421/2020\|EPI_ISL_741371\|20 | ···················· | ·A·················· | ···················· | ···················· | ···················· | ··A················· |
| Z | 5811 | hCoV-19/England/MILK-B3D44F/2020\|EPI_ISL_741372\|20 | ···················· | ···················· | ···················· | ···················· | ···················· | ···················· |
| Z | 5812 | hCoV-19/England/MILK-B3D45E/2020\|EPI_ISL_741373\|20 | ···················· | ···················· | ···················· | ···················· | ···················· | ···················· |
| Z | 5813 | hCoV-19/England/MILK-B3D99B/2020\|EPI_ISL_741374\|20 | ···················· | ···················· | ···················· | ···················· | ···················· | ···················· |
| Z | 5814 | hCoV-19/England/MILK-B3D9AA/2020\|EPI_ISL_741375\|20 | ···················· | ···················· | ···················· | ···················· | ······T············· | ···················· |
| Z | 5815 | hCoV-19/England/MILK-B3D9C8/2020\|EPI_ISL_741376\|20 | ···················· | ···················· | ···················· | ···················· | ···················· | ···················· |
| Z | 5816 | hCoV-19/England/MILK-B3D9E6/2020\|EPI_ISL_741377\|20 | ···················· | ·A·················· | ···················· | ···················· | ···················· | ··A················· |
| Z | 5817 | hCoV-19/England/MILK-B3D9F5/2020\|EPI_ISL_741378\|20 | ···················· | ···················· | ···················· | ···················· | ···················· | ···················· |
| Z | 5818 | hCoV-19/England/MILK-B3DA4D/2020\|EPI_ISL_741380\|20 | ···················· | ···················· | ···················· | ···················· | ···················· | ···················· |
| Z | 5819 | hCoV-19/England/MILK-B3DA5C/2020\|EPI_ISL_741381\|20 | ···················· | ···················· | ···················· | ···················· | ···················· | ···················· |
| Z | 5820 | hCoV-19/England/MILK-B3DA89/2020\|EPI_ISL_741382\|20 | ···················· | ···················· | ···················· | ···················· | ···················· | ···················· |
| Z | 5821 | hCoV-19/England/MILK-B3DA98/2020\|EPI_ISL_741383\|20 | ···················· | ···················· | ···················· | ···················· | ···················· | ···················· |
| Z | 5822 | hCoV-19/England/MILK-B3DAB6/2020\|EPI_ISL_741384\|20 | ···················· | ···················· | ···················· | ···················· | ···················· | ···················· |
| Z | 5823 | hCoV-19/England/MILK-B3DAC5/2020\|EPI_ISL_741385\|20 | ···················· | ···················· | ···················· | ···················· | ···················· | ···················· |
| Z | 5824 | hCoV-19/England/MILK-B3DAD4/2020\|EPI_ISL_741386\|20 | ···················· | ···················· | ···················· | ···················· | ···················· | ···················· |
| Z | 5825 | hCoV-19/England/MILK-B3DAE3/2020\|EPI_ISL_741387\|20 | ···················· | ···················· | ···················· | ···················· | ···················· | ···················· |
| Z | 5826 | hCoV-19/England/MILK-B3DAF2/2020\|EPI_ISL_741388\|20 | ···················· | ···A·····T·········· | ···················· | ···················· | ···················· | ···················· |
| Z | 5827 | hCoV-19/England/MILK-B3DB1D/2020\|EPI_ISL_741389\|20 | ···················· | ···················· | ···················· | ···················· | ···················· | ···················· |
| Z | 5828 | hCoV-19/England/MILK-B3DB2C/2020\|EPI_ISL_741390\|20 | ···················· | ···················· | ···················· | ···················· | ······T············· | ···················· |
| Z | 5829 | hCoV-19/England/MILK-B3DB4A/2020\|EPI_ISL_741391\|20 | ···················· | ·A·················· | ···················· | ···················· | ···················· | ··A················· |
| Z | 5830 | hCoV-19/England/MILK-B3DB59/2020\|EPI_ISL_741392\|20 | ···················· | ···················· | ···················· | ···················· | ···················· | ···················· |
| Z | 5831 | hCoV-19/England/MILK-B3DB68/2020\|EPI_ISL_741393\|20 | ···················· | ···················· | ···················· | ···················· | ···················· | ···················· |
| Z | 5832 | hCoV-19/England/MILK-B3DB77/2020\|EPI_ISL_741394\|20 | ···················· | ···················· | ···················· | ···················· | ···················· | ···················· |
| Z | 5833 | hCoV-19/England/MILK-B3DDAE/2020\|EPI_ISL_741395\|20 | ···················· | ···················· | ···················· | ···················· | ···················· | ···················· |
| Z | 5834 | hCoV-19/England/MILK-B3DDCC/2020\|EPI_ISL_741396\|20 | ···················· | ···················· | ···················· | ···················· | ···················· | ···················· |
| Z | 5835 | hCoV-19/England/MILK-B3DDDB/2020\|EPI_ISL_741397\|20 | ···················· | ·A·················· | ···················· | ···················· | ···················· | ··A················· |
| Z | 5836 | hCoV-19/England/MILK-B3DDF9/2020\|EPI_ISL_741398\|20 | ···················· | ···················· | ···················· | ···················· | ···················· | ···················· |
| Z | 5837 | hCoV-19/England/MILK-B3DE14/2020\|EPI_ISL_741399\|20 | ···················· | ···················· | ···················· | ···················· | ···················· | ···················· |
| Z | 5838 | hCoV-19/Wales/PHWC-49CAF8/2020\|EPI_ISL_742645\|2020 | ···················· | ···················· | ···················· | ···················· | ···················· | ···················· |
| Z | 5839 | hCoV-19/Wales/PHWC-49CB04/2020\|EPI_ISL_742646\|2020 | ···················· | ···················· | ···················· | ···················· | ···················· | ···················· |
| Z | 5840 | hCoV-19/Wales/PHWC-49CB13/2020\|EPI_ISL_742647\|2020 | ···················· | ···················· | ···················· | ···················· | ···················· | ···················· |
| Z | 5841 | hCoV-19/Wales/PHWC-49CB31/2020\|EPI_ISL_742648\|2020 | ···················· | ···················· | ···················· | ···················· | ···················· | ···················· |
| Z | 5842 | hCoV-19/Wales/PHWC-49CB40/2020\|EPI_ISL_742649\|2020 | ···················· | ···················· | ···················· | ···················· | ···················· | ···················· |
| Z | 5843 | hCoV-19/Wales/PHWC-49CB6E/2020\|EPI_ISL_742651\|2020 | ···················· | ···················· | ···················· | ···················· | ···················· | ···················· |
| Z | 5844 | hCoV-19/Wales/PHWC-49CB7D/2020\|EPI_ISL_742652\|2020 | ···················· | ···················· | ···················· | ···················· | ···················· | ···················· |
| Z | 5845 | hCoV-19/Wales/PHWC-49CB8C/2020\|EPI_ISL_742653\|2020 | ···················· | ···················· | ···················· | ···················· | ···················· | ···················· |
| Z | 5847 | hCoV-19/Wales/PHWC-49CBAA/2020\|EPI_ISL_742655\|2020 | ···················· | ···················· | ···················· | ···················· | ···················· | ···················· |

| | | | | | | | |
|---|---|---|---|---|---|---|---|
| Z | 5848 hCoV-19/Wales/PHWC-49CBB9/2020\|EPI_ISL_742656\|2020 | ···················· | ···················· | ···················· | ···················· | ···················· | ···················· |
| Z | 5849 hCoV-19/Wales/PHWC-49CBC8/2020\|EPI_ISL_742657\|2020 | ···················· | ···················· | ···················· | ···················· | ···················· | ···················· |
| Z | 5850 hCoV-19/Wales/PHWC-49CBD7/2020\|EPI_ISL_742658\|2020 | ···················· | ···················· | ···················· | ···················· | ···················· | ···················· |
| Z | 5852 hCoV-19/Wales/PHWC-49CBF5/2020\|EPI_ISL_742660\|2020 | ···················· | ···················· | ···················· | ···················· | ···················· | ···················· |
| Z | 5853 hCoV-19/Wales/PHWC-49CC01/2020\|EPI_ISL_742661\|2020 | ···················· | ···················· | ···················· | ···················· | ···················· | ···················· |
| Z | 5854 hCoV-19/Wales/PHWC-49CC10/2020\|EPI_ISL_742662\|2020 | ···················· | ···················· | ···················· | ···················· | ···················· | ···················· |
| Z | 5855 hCoV-19/Wales/PHWC-49CC2F/2020\|EPI_ISL_742663\|2020 | ···················· | ···················· | ···················· | ···················· | ···················· | ···················· |
| Z | 5856 hCoV-19/Wales/PHWC-49CC3E/2020\|EPI_ISL_742664\|2020 | ···················· | ···················· | ···················· | ···················· | ···················· | ···················· |
| Z | 5857 hCoV-19/Wales/PHWC-49CC5C/2020\|EPI_ISL_742666\|2020 | ···················· | ···················· | ···················· | ···················· | ···················· | ···················· |
| Z | 5858 hCoV-19/Wales/PHWC-49CC6B/2020\|EPI_ISL_742667\|2020 | ···················· | ···················· | ···················· | ···················· | ···················· | ···················· |
| Z | 5859 hCoV-19/Wales/PHWC-49CC7A/2020\|EPI_ISL_742668\|2020 | ···················· | ···················· | ···················· | ···················· | ···················· | ···················· |
| Z | 5860 hCoV-19/Wales/PHWC-49CC89/2020\|EPI_ISL_742669\|2020 | ···················· | ···················· | ···················· | ···················· | ···················· | ···················· |
| Z | 5861 hCoV-19/Wales/PHWC-49CDD1/2020\|EPI_ISL_742689\|2020 | ···················· | ···················· | ···················· | ···················· | ···················· | ···················· |
| Z | 5862 hCoV-19/Wales/PHWC-49CDFF/2020\|EPI_ISL_742691\|2020 | ···················· | ···················· | ···················· | ···················· | ···················· | ···················· |
| Z | 5863 hCoV-19/Wales/PHWC-49CE0B/2020\|EPI_ISL_742692\|2020 | ···················· | ···················· | ···················· | ···················· | ···················· | ···················· |
| Z | 5864 hCoV-19/Wales/PHWC-49CE1A/2020\|EPI_ISL_742693\|2020 | ···················· | ···················· | ···················· | ···················· | ···················· | ···················· |
| Z | 5865 hCoV-19/Wales/PHWC-49CE29/2020\|EPI_ISL_742694\|2020 | ···················· | ···················· | ···················· | ···················· | ···················· | ···················· |
| Z | 5867 hCoV-19/Wales/PHWC-49CE47/2020\|EPI_ISL_742696\|2020 | ···················· | ···················· | ···················· | ···················· | ···················· | ···················· |
| Z | 5868 hCoV-19/Wales/PHWC-49CE56/2020\|EPI_ISL_742697\|2020 | ···················· | ···················· | ···················· | ···················· | ···················· | ···················· |
| Z | 5869 hCoV-19/Wales/PHWC-49CE65/2020\|EPI_ISL_742698\|2020 | ···················· | ···················· | ···················· | ···················· | ···················· | ···················· |
| Z | 5870 hCoV-19/Wales/PHWC-49CE74/2020\|EPI_ISL_742699\|2020 | ···················· | ···················· | ···················· | ···················· | ···················· | ···················· |
| Z | 5871 hCoV-19/Wales/PHWC-4A2AC2/2020\|EPI_ISL_743976\|2020 | ···················· | ···················· | ···················· | ···················· | ···················· | ···················· |
| Z | 5872 hCoV-19/Wales/PHWC-4A2AE0/2020\|EPI_ISL_743978\|2020 | ···················· | ···················· | ···················· | ···················· | ···················· | ···················· |
| Z | 5873 hCoV-19/Wales/PHWC-4A2B29/2020\|EPI_ISL_743982\|2020 | ···················· | ···················· | ···················· | ···················· | ···················· | ···················· |
| Z | 5874 hCoV-19/Wales/PHWC-4A2B47/2020\|EPI_ISL_743983\|2020 | ···················· | ···················· | ···················· | ···················· | ···················· | ···················· |
| Z | 5875 hCoV-19/Wales/PHWC-4A2B56/2020\|EPI_ISL_743984\|2020 | ···················· | ···················· | ···················· | ···················· | ···················· | ···················· |
| Z | 5876 hCoV-19/Wales/PHWC-4A2B65/2020\|EPI_ISL_743985\|2020 | ···················· | ···················· | ···················· | ···················· | ···················· | ···················· |
| Z | 5877 hCoV-19/Wales/PHWC-4A2B74/2020\|EPI_ISL_743986\|2020 | ···················· | ···················· | ···················· | ···················· | ···················· | ···················· |
| Z | 5878 hCoV-19/Wales/PHWC-4A2BA1/2020\|EPI_ISL_743988\|2020 | ···················· | ···················· | ···················· | ···················· | ···················· | ···················· |
| Z | 5879 hCoV-19/Wales/PHWC-4A2BB0/2020\|EPI_ISL_743989\|2020 | ···················· | ···················· | ···················· | ···················· | ···················· | ···················· |
| Z | 5880 hCoV-19/Wales/PHWC-4A2BCF/2020\|EPI_ISL_743990\|2020 | ···················· | ···················· | ···················· | ···················· | ···················· | ···················· |
| Z | 5881 hCoV-19/Wales/PHWC-4A2BDE/2020\|EPI_ISL_743991\|2020 | ···················· | ···················· | ···················· | ···················· | ···················· | ···················· |
| Z | 5882 hCoV-19/Wales/PHWC-4A2BED/2020\|EPI_ISL_743992\|2020 | ···················· | ···················· | ···················· | ···················· | ···················· | ···················· |
| Z | 5883 hCoV-19/Wales/PHWC-4A2BFC/2020\|EPI_ISL_743993\|2020 | ···················· | ···················· | ···················· | ···················· | ···················· | ···················· |
| Z | 5884 hCoV-19/Wales/PHWC-4A2C08/2020\|EPI_ISL_743994\|2020 | ···················· | ···················· | ···················· | ···················· | ···················· | ···················· |
| Z | 5886 hCoV-19/Wales/PHWC-4A2C26/2020\|EPI_ISL_743996\|2020 | ···················· | ···················· | ···················· | ···················· | ···················· | ···················· |
| Z | 5887 hCoV-19/Wales/PHWC-4A2C44/2020\|EPI_ISL_743997\|2020 | ···················· | ···················· | ···················· | ···················· | ···················· | ···················· |
| Z | 5888 hCoV-19/Wales/PHWC-4A2C53/2020\|EPI_ISL_743998\|2020 | ···················· | ···················· | ···················· | ···················· | ···················· | ···················· |
| Z | 5889 hCoV-19/USA/CA-CZB-15961/2020\|EPI_ISL_739087\|2020- | ···················· | ···················· | ···················· | ···················· | ···················· | ···················· |
| Z | 5890 hCoV-19/USA/CA-CZB-14296/2020\|EPI_ISL_739088\|2020- | ···················· | ···················· | ···················· | ···················· | ···················· | ···················· |
| Z | 5892 hCoV-19/USA/CA-CZB-14016/2020\|EPI_ISL_739100\|2020- | ···················· | ···················· | ···················· | ···················· | ···················· | ···················· |
| Z | 5893 hCoV-19/USA/CA-CZB-14954/2020\|EPI_ISL_739101\|2020- | ···················· | ···················· | ···················· | ···················· | ···················· | ···················· |
| Z | 5894 hCoV-19/USA/CA-CZB-15495/2020\|EPI_ISL_739102\|2020- | ···················· | ···················· | ···················· | ···················· | ···················· | ···················· |
| Z | 5895 hCoV-19/USA/CA-CZB-15000/2020\|EPI_ISL_739103\|2020- | ···················· | ···················· | ···················· | ···················· | ···················· | ···················· |
| Z | 5896 hCoV-19/USA/CA-CZB-15956/2020\|EPI_ISL_739104\|2020- | ···················· | ·A·················· | ···················· | ···················· | ···················· | ···················· |
| Z | 5897 hCoV-19/USA/CA-CZB-14237/2020\|EPI_ISL_739105\|2020- | ···················· | ···················· | ···················· | ···················· | ···················· | ···················· |
| Z | 5898 hCoV-19/USA/CA-CZB-14846/2020\|EPI_ISL_739106\|2020- | ···················· | ···················· | ···················· | ···················· | ···················· | ···················· |
| Z | 5899 hCoV-19/USA/CA-CZB-13699/2020\|EPI_ISL_739107\|2020- | ···················· | ···················· | ···················· | ···················· | ···················· | ···················· |
| Z | 5900 hCoV-19/USA/CA-CZB-14201/2020\|EPI_ISL_739108\|2020- | ···················· | ·R·················· | ···················· | ···················· | ···················· | ···················· |
| Z | 5901 hCoV-19/USA/CA-CZB-13834/2020\|EPI_ISL_739109\|2020- | ···················· | ···················· | ···················· | ···················· | ···················· | ···················· |
| Z | 5903 hCoV-19/USA/CA-CZB-14060/2020\|EPI_ISL_739111\|2020- | ···················· | ···················· | ···················· | ···················· | ···················· | ···················· |
| Z | 5904 hCoV-19/USA/CA-CZB-14803/2020\|EPI_ISL_739112\|2020- | ···················· | ···················· | ···················· | ···················· | ···················· | ···················· |
| Z | 5906 hCoV-19/USA/CA-CZB-14818/2020\|EPI_ISL_739114\|2020- | ···················· | ···················· | ···················· | ···················· | ···················· | ···················· |
| Z | 5907 hCoV-19/USA/CA-CZB-13807/2020\|EPI_ISL_739115\|2020- | ···················· | ···················· | ···················· | ···················· | ···················· | ···················· |
| Z | 5908 hCoV-19/USA/CA-CZB-15426/2020\|EPI_ISL_739116\|2020- | ···················· | ·A·················· | ···················· | ···················· | ···················· | ···R················ |
| Z | 5909 hCoV-19/USA/CA-CZB-15291/2020\|EPI_ISL_739117\|2020- | ···················· | ·A·················· | ···················· | ···················· | ···················· | ···················· |
| Z | 5911 hCoV-19/USA/CA-CZB-14712/2020\|EPI_ISL_739119\|2020- | ·······T············ | ···················· | ···················· | ···················· | ···················· | ···················· |
| Z | 5912 hCoV-19/USA/CA-CZB-14082/2020\|EPI_ISL_739140\|2020- | ···················· | ···················· | ···················· | ···················· | ···················· | ···················· |
| Z | 5913 hCoV-19/USA/CA-CZB-15966/2020\|EPI_ISL_739141\|2020- | ···················· | ·A·················· | ···················· | ···················· | ···················· | ···················· |
| Z | 5914 hCoV-19/USA/CA-CZB-13905/2020\|EPI_ISL_739150\|2020- | ···················· | ·A·················· | ···················· | ···················· | ···················· | ···················· |
| Z | 5915 hCoV-19/USA/CA-CZB-14200/2020\|EPI_ISL_739151\|2020- | ···················· | ···················· | ···················· | ···················· | ···················· | ···················· |
| Z | 5917 hCoV-19/USA/CA-CZB-15311/2020\|EPI_ISL_739153\|2020- | ···················· | ·A·················· | ···················· | ···················· | ···················· | ···················· |
| Z | 5918 hCoV-19/USA/CA-CZB-14936/2020\|EPI_ISL_739154\|2020- | ···················· | ···················· | ···················· | ···················· | ···················· | ···················· |
| Z | 5919 hCoV-19/USA/CA-CZB-14853/2020\|EPI_ISL_739155\|2020- | ···················· | ···················· | ···················· | ···················· | ···················· | ···················· |
| Z | 5920 hCoV-19/USA/CA-CZB-14209/2020\|EPI_ISL_739156\|2020- | ···················· | ·············A······ | ···················· | ···················· | ···················· | ···················· |
| Z | 5921 hCoV-19/USA/CA-CZB-13673/2020\|EPI_ISL_739157\|2020- | ···················· | ·A·················· | ···················· | ···················· | ···················· | ···················· |
| Z | 5922 hCoV-19/USA/CA-CZB-13864/2020\|EPI_ISL_739158\|2020- | ···················· | ···················· | ···················· | ···················· | ···················· | ···················· |
| Z | 5923 hCoV-19/USA/CA-CZB-14861/2020\|EPI_ISL_739159\|2020- | ···················· | ···················· | ···················· | ···················· | ···················· | ···················· |
| Z | 5924 hCoV-19/Luxembourg/LNS2335071/2020\|EPI_ISL_740100\| | ···········T········ | ···················· | ···················· | ···················· | ···················· | ···················· |
| Z | 5925 hCoV-19/Luxembourg/LNS2339656/2020\|EPI_ISL_740101\| | ···················· | ···················· | ···················· | ···················· | ···················· | ···················· |
| Z | 5926 hCoV-19/Luxembourg/LNS2346680/2020\|EPI_ISL_740102\| | ···················· | ···················· | ···················· | ···················· | ···················· | ···················· |
| Z | 5927 hCoV-19/Luxembourg/LNS2350135/2020\|EPI_ISL_740103\| | ···················· | ···················· | ···················· | ···················· | ···················· | ···················· |
| Z | 5928 hCoV-19/Luxembourg/LNS2493379/2020\|EPI_ISL_740130\| | ···················· | ···················· | ···················· | ···················· | ···················· | ···················· |
| Z | 5929 hCoV-19/Luxembourg/LNS2500324/2020\|EPI_ISL_740131\| | ···················· | ···················· | ···················· | ···················· | ···················· | ···················· |
| Z | 5930 hCoV-19/Luxembourg/LNS2509324/2020\|EPI_ISL_740132\| | ···················· | ···················· | ···················· | ···················· | ···················· | ···················· |
| Z | 5931 hCoV-19/Luxembourg/LNS2510787/2020\|EPI_ISL_740133\| | ···················· | ···················· | ···················· | ···················· | ···················· | ···················· |
| Z | 5932 hCoV-19/Luxembourg/LNS2529691/2020\|EPI_ISL_740137\| | ···················· | ···················· | ···················· | ···················· | ···················· | ···················· |
| Z | 5933 hCoV-19/Luxembourg/LNS2540162/2020\|EPI_ISL_740139\| | ···················· | ···················· | ···················· | ···················· | ···················· | ···················· |
| Z | 5934 hCoV-19/Luxembourg/LNS2545613/2020\|EPI_ISL_740140\| | ···················· | ···················· | ···················· | ···················· | ···················· | ···················· |
| Z | 5935 hCoV-19/Luxembourg/LNS2552311/2020\|EPI_ISL_740141\| | ···················· | ···················· | ···················· | ···················· | ···················· | ···················· |

| | | | | | | | | | |
|---|---|---|---|---|---|---|---|---|---|
| Z | 5936 | hCoV-19/Luxembourg/LNS2552420/2020\|EPI_ISL_740142\| | ···················· | ···················· | ···················· | ···················· | ···················· | ···················· | |
| Z | 5937 | hCoV-19/Luxembourg/LNS2555815/2020\|EPI_ISL_740143\| | ···················· | ···················· | ···················· | ···················· | ···················· | ···················· | |
| Z | 5938 | hCoV-19/Luxembourg/LNS2558747/2020\|EPI_ISL_740144\| | ···················· | ···················· | ···················· | ···················· | ···················· | ···················· | |
| Z | 5939 | hCoV-19/Luxembourg/LNS2567494/2020\|EPI_ISL_740145\| | ···················· | ···················· | ···················· | ···················· | ···················· | ···················· | |
| Z | 5940 | hCoV-19/Luxembourg/LNS2580720/2020\|EPI_ISL_740146\| | ···················· | ···················· | ···················· | ···················· | ···················· | ···················· | |
| Z | 5941 | hCoV-19/Luxembourg/LNS2581393/2020\|EPI_ISL_740147\| | ···················· | ···················· | ···················· | ···················· | ···················· | ···················· | |
| Z | 5942 | hCoV-19/England/MILK-B3DE41/2020\|EPI_ISL_741401\|20 | ···················· | ···················· | ···················· | ···················· | ···················· | ···················· | |
| Z | 5943 | hCoV-19/England/MILK-B3DE50/2020\|EPI_ISL_741402\|20 | ···················· | ···················· | ···················· | ···················· | ···················· | ···················· | |
| Z | 5944 | hCoV-19/England/MILK-B3DE6F/2020\|EPI_ISL_741403\|20 | ···················· | ···················· | ···················· | ···················· | ···················· | ···················· | |
| Z | 5945 | hCoV-19/England/MILK-B3DE8D/2020\|EPI_ISL_741404\|20 | ···················· | ·A·················· | ···················· | ···················· | ···················· | ···················· | |
| Z | 5946 | hCoV-19/England/MILK-B3DE9C/2020\|EPI_ISL_741405\|20 | ···················· | ···················· | ···················· | ···················· | ···················· | ···················· | |
| Z | 5947 | hCoV-19/England/MILK-B3DEBA/2020\|EPI_ISL_741406\|20 | ···················· | ···················· | ···················· | ···················· | ···················· | ···················· | |
| Z | 5948 | hCoV-19/England/MILK-B3DEC9/2020\|EPI_ISL_741407\|20 | ···················· | ···················· | ···················· | ···················· | ···················· | ···················· | |
| Z | 5949 | hCoV-19/England/MILK-B3DEF6/2020\|EPI_ISL_741408\|20 | ···················· | ···················· | ···················· | ···················· | ···················· | ···················· | |
| Z | 5950 | hCoV-19/England/MILK-B3DF20/2020\|EPI_ISL_741409\|20 | ···················· | ···················· | ···················· | ···················· | ···················· | ···················· | |
| Z | 5951 | hCoV-19/England/MILK-B3DF4E/2020\|EPI_ISL_741410\|20 | ···················· | ···················· | ···················· | ···················· | ···················· | ···················· | |
| Z | 5952 | hCoV-19/England/MILK-B3DF5D/2020\|EPI_ISL_741411\|20 | ···················· | ···················· | ···················· | ···················· | ···················· | ···················· | |
| Z | 5953 | hCoV-19/England/MILK-B3DF6C/2020\|EPI_ISL_741412\|20 | ···················· | ···················· | ···················· | ···················· | ···················· | ···················· | |
| Z | 5954 | hCoV-19/England/MILK-B3DF7B/2020\|EPI_ISL_741413\|20 | ···················· | ·A·················· | ···················· | ···················· | ···················· | ··A················· | |
| Z | 5955 | hCoV-19/England/MILK-B3DF8A/2020\|EPI_ISL_741414\|20 | ···················· | ···················· | ···················· | ···················· | ···················· | ···················· | |
| Z | 5956 | hCoV-19/England/MILK-B3DF99/2020\|EPI_ISL_741415\|20 | ···················· | ···················· | ···················· | ···················· | ···················· | ···················· | |
| Z | 5957 | hCoV-19/England/MILK-B3DFA8/2020\|EPI_ISL_741416\|20 | ···················· | ···················· | ···················· | ···················· | ···················· | ···················· | |
| Z | 5958 | hCoV-19/England/MILK-B3DFB7/2020\|EPI_ISL_741417\|20 | ···················· | ···················· | ···················· | ···················· | ···················· | ···················· | |
| Z | 5959 | hCoV-19/England/MILK-B3E280/2020\|EPI_ISL_741419\|20 | ···················· | ···················· | ···················· | ···················· | ···················· | ···················· | |
| Z | 5960 | hCoV-19/England/MILK-B3E314/2020\|EPI_ISL_741420\|20 | ···················· | ···················· | ···················· | ···················· | ···················· | ···················· | |
| Z | 5961 | hCoV-19/England/NORT-1B62FF2/2020\|EPI_ISL_741421\|2 | ···················· | ···················· | ···················· | ···················· | ···················· | ···················· | |
| Z | 5962 | hCoV-19/England/NORT-1B630D0/2020\|EPI_ISL_741423\|2 | ···················· | ···················· | ···················· | ···················· | ···················· | ···················· | |
| Z | 5963 | hCoV-19/England/NORT-1B63252/2020\|EPI_ISL_741426\|2 | ···················· | ···················· | ···················· | ···················· | ···················· | ···················· | |
| Z | 5964 | hCoV-19/England/NORT-1B6329E/2020\|EPI_ISL_741427\|2 | ···················· | ···················· | ···················· | ···················· | ···················· | ···················· | |
| Z | 5965 | hCoV-19/England/NORT-1B632AD/2020\|EPI_ISL_741428\|2 | ···················· | ···················· | ···················· | ···················· | ···················· | ···················· | |
| Z | 5966 | hCoV-19/England/NORT-1B79602/2020\|EPI_ISL_741429\|2 | ···················· | ···················· | ···················· | ···················· | ······T············· | ···················· | |
| Z | 5967 | hCoV-19/England/NORT-1B7968A/2020\|EPI_ISL_741430\|2 | ···················· | ···················· | ···················· | ···················· | ···················· | ···················· | |
| Z | 5968 | hCoV-19/England/NORT-1B7970F/2020\|EPI_ISL_741432\|2 | ···················· | ···················· | ···················· | ···················· | ···················· | ···················· | |
| Z | 5969 | hCoV-19/England/NORT-1B79769/2020\|EPI_ISL_741433\|2 | ···················· | ···················· | ···················· | ···················· | ···················· | ···················· | |
| Z | 5970 | hCoV-19/England/NORW-21FAA4/2020\|EPI_ISL_741460\|20 | ···················· | ···················· | ···················· | ···················· | ···················· | ···················· | |
| Z | 5971 | hCoV-19/England/NORW-21FB65/2020\|EPI_ISL_741464\|20 | ···················· | ···················· | ···················· | ···················· | ···················· | ···················· | |
| Z | 5972 | hCoV-19/England/NORW-21FB83/2020\|EPI_ISL_741465\|20 | ···················· | ···················· | ···················· | ···················· | ···················· | ···················· | |
| Z | 5973 | hCoV-19/England/NORW-21FC26/2020\|EPI_ISL_741469\|20 | ···················· | ·A·················· | ···················· | ···················· | ···················· | ··A················· | |
| Z | 5974 | hCoV-19/England/NORW-21FC53/2020\|EPI_ISL_741470\|20 | ···················· | ···················· | ···················· | ···················· | ···················· | ···················· | |
| Z | 5975 | hCoV-19/England/NORW-21FC71/2020\|EPI_ISL_741471\|20 | ···················· | ···················· | ···················· | ···················· | ···················· | ···················· | |
| Z | 5976 | hCoV-19/England/NORW-21FC80/2020\|EPI_ISL_741472\|20 | ···················· | ···················· | ···················· | ···················· | ···················· | ···················· | |
| Z | 5977 | hCoV-19/England/NORW-21FCBD/2020\|EPI_ISL_741473\|20 | ···················· | ···················· | ···················· | ···················· | ···················· | ···················· | |
| Z | 5978 | hCoV-19/England/NORW-21FCDB/2020\|EPI_ISL_741474\|20 | ···················· | ···················· | ···················· | ···················· | ···················· | ···················· | |
| Z | 5979 | hCoV-19/England/NORW-21FCF9/2020\|EPI_ISL_741475\|20 | ···················· | ···················· | ···················· | ···················· | ···················· | ···················· | |
| Z | 5980 | hCoV-19/England/NORW-21FD32/2020\|EPI_ISL_741476\|20 | ···················· | ···················· | ···················· | ···················· | ···················· | ···················· | |
| Z | 5981 | hCoV-19/England/NORW-21FD50/2020\|EPI_ISL_741477\|20 | ···················· | ···················· | ···················· | ···················· | ···················· | ···················· | |
| Z | 5982 | hCoV-19/England/NORW-21FD7E/2020\|EPI_ISL_741478\|20 | ···················· | ···················· | ···················· | ···················· | ···················· | ···················· | |
| Z | 5983 | hCoV-19/Wales/PHWC-49CE83/2020\|EPI_ISL_742700\|2020 | ···················· | ···················· | ···················· | ···················· | ···················· | ···················· | |
| Z | 5984 | hCoV-19/Wales/PHWC-49CEA1/2020\|EPI_ISL_742701\|2020 | ···················· | ···················· | ···················· | ···················· | ···················· | ···················· | |
| Z | 5985 | hCoV-19/Wales/PHWC-49CEB0/2020\|EPI_ISL_742702\|2020 | ···················· | ···················· | ···················· | ···················· | ···················· | ···················· | |
| Z | 5986 | hCoV-19/Wales/PHWC-49CEDE/2020\|EPI_ISL_742704\|2020 | ···················· | ···················· | ···················· | ···················· | ···················· | ···················· | |
| Z | 5987 | hCoV-19/Wales/PHWC-49CEFC/2020\|EPI_ISL_742705\|2020 | ···················· | ···················· | ···················· | ···················· | ···················· | ···················· | |
| Z | 5988 | hCoV-19/Wales/PHWC-49CF08/2020\|EPI_ISL_742706\|2020 | ···················· | ···················· | ···················· | ···················· | ···················· | ···················· | |
| Z | 5990 | hCoV-19/Wales/PHWC-49CF26/2020\|EPI_ISL_742708\|2020 | ···················· | ···················· | ···················· | ···················· | ···················· | ···················· | |
| Z | 5991 | hCoV-19/Wales/PHWC-49CF35/2020\|EPI_ISL_742709\|2020 | ···················· | ···················· | ···················· | ···················· | ···················· | ···················· | |
| Z | 5992 | hCoV-19/Wales/PHWC-49CF62/2020\|EPI_ISL_742710\|2020 | ···················· | ···················· | ···················· | ···················· | ···················· | ···················· | |
| Z | 5993 | hCoV-19/Wales/PHWC-49CF80/2020\|EPI_ISL_742712\|2020 | ···················· | ···················· | ···················· | ···················· | ···················· | ···················· | |
| Z | 5994 | hCoV-19/Wales/PHWC-49CF9F/2020\|EPI_ISL_742713\|2020 | ···················· | ···················· | ···················· | ···················· | ···················· | ···················· | |
| Z | 5995 | hCoV-19/Wales/PHWC-49CFAE/2020\|EPI_ISL_742714\|2020 | ···················· | ···················· | ···················· | ···················· | ···················· | ···················· | |
| Z | 5996 | hCoV-19/Wales/PHWC-49CFBD/2020\|EPI_ISL_742715\|2020 | ···················· | ···················· | ···················· | ···················· | ···················· | ···················· | |
| Z | 5997 | hCoV-19/Wales/PHWC-49CFCC/2020\|EPI_ISL_742716\|2020 | ···················· | ···················· | ···················· | ···················· | NNN················· | ···················· | 1 |
| Z | 5998 | hCoV-19/Wales/PHWC-49CFDB/2020\|EPI_ISL_742717\|2020 | ···················· | ···················· | ···················· | ···················· | ···················· | ···················· | |
| Z | 5999 | hCoV-19/Wales/PHWC-49CFEA/2020\|EPI_ISL_742718\|2020 | ···················· | ···················· | ···················· | ···················· | ···················· | ···················· | |
| Z | 6000 | hCoV-19/Wales/PHWC-49CFF9/2020\|EPI_ISL_742719\|2020 | ···················· | ···················· | ···················· | ···················· | ···················· | ···················· | |
| Z | 6001 | hCoV-19/Wales/PHWC-49D013/2020\|EPI_ISL_742721\|2020 | ···················· | ···················· | ···················· | ···················· | ···················· | ···················· | |
| Z | 6002 | hCoV-19/Wales/PHWC-49D022/2020\|EPI_ISL_742722\|2020 | ···················· | ···················· | ···················· | ···················· | ···················· | ···················· | |
| Z | 6003 | hCoV-19/Wales/PHWC-49D031/2020\|EPI_ISL_742723\|2020 | ···················· | ···················· | ···················· | ···················· | ···················· | ···················· | |
| Z | 6004 | hCoV-19/Wales/PHWC-49D040/2020\|EPI_ISL_742724\|2020 | ···················· | ···················· | ···················· | ···················· | ···················· | ···················· | |
| Z | 6005 | hCoV-19/Wales/PHWC-49D05F/2020\|EPI_ISL_742725\|2020 | ···················· | ···················· | ···················· | ···················· | ···················· | ···················· | |
| Z | 6006 | hCoV-19/Wales/PHWC-49D06E/2020\|EPI_ISL_742726\|2020 | ···················· | ···················· | ···················· | ···················· | ···················· | ···················· | |
| Z | 6007 | hCoV-19/Wales/PHWC-49D07D/2020\|EPI_ISL_742727\|2020 | ···················· | ···················· | ···················· | ···················· | ···················· | ···················· | |
| Z | 6008 | hCoV-19/Wales/PHWC-49D08C/2020\|EPI_ISL_742728\|2020 | ···················· | ···················· | ···················· | ···················· | ···················· | ···················· | |
| Z | 6009 | hCoV-19/Wales/PHWC-49D0B9/2020\|EPI_ISL_742730\|2020 | ···················· | ···················· | ···················· | ···················· | ···················· | ···················· | |
| Z | 6011 | hCoV-19/Wales/PHWC-49D0D7/2020\|EPI_ISL_742732\|2020 | ···················· | ···················· | ···················· | ···················· | ···················· | ···················· | |
| Z | 6013 | hCoV-19/Wales/PHWC-49D0F5/2020\|EPI_ISL_742734\|2020 | ···················· | ···················· | ···················· | ···················· | ···················· | ···················· | |
| Z | 6014 | hCoV-19/Wales/PHWC-49D101/2020\|EPI_ISL_742735\|2020 | ···················· | ···················· | ···················· | ···················· | ···················· | ···················· | |
| Z | 6015 | hCoV-19/Wales/PHWC-49D110/2020\|EPI_ISL_742736\|2020 | ···················· | ···················· | ···················· | ···················· | ···················· | ···················· | |
| Z | 6016 | hCoV-19/Wales/PHWC-49D12F/2020\|EPI_ISL_742737\|2020 | ···················· | ···················· | ···················· | ···················· | ···················· | ···················· | |
| Z | 6017 | hCoV-19/Wales/PHWC-49D13E/2020\|EPI_ISL_742738\|2020 | ···················· | ···················· | ···················· | ···················· | ···················· | ···················· | |
| Z | 6018 | hCoV-19/Wales/PHWC-49D14D/2020\|EPI_ISL_742739\|2020 | ···················· | ···················· | ···················· | ···················· | ···················· | ···················· | |

| | | | | | | | |
|---|---|---|---|---|---|---|---|
| Z | 6019 hCoV-19/Wales/PHWC-49D189/2020\|EPI_ISL_742742\|2020 | ···················· | ···················· | ····················· | ···················· | ···················· | ···················· |
| Z | 6020 hCoV-19/Wales/PHWC-49D1A7/2020\|EPI_ISL_742744\|2020 | ···················· | ···················· | ····················· | ···················· | ···················· | ···················· |
| Z | 6021 hCoV-19/Wales/PHWC-49D1B6/2020\|EPI_ISL_742745\|2020 | ···················· | ···················· | ····················· | ···················· | ···················· | ···················· |
| Z | 6023 hCoV-19/Wales/PHWC-49D1D4/2020\|EPI_ISL_742747\|2020 | ···················· | ···················· | ····················· | ···················· | ···················· | ···················· |
| Z | 6024 hCoV-19/Wales/PHWC-49D1E3/2020\|EPI_ISL_742748\|2020 | ···················· | ···················· | ····················· | ···················· | ···················· | ···················· |
| Z | 6025 hCoV-19/Wales/PHWC-49D1F2/2020\|EPI_ISL_742749\|2020 | ···················· | ···················· | ····················· | ···················· | ···················· | ···················· |
| Z | 6026 hCoV-19/Wales/PHWC-49D20E/2020\|EPI_ISL_742750\|2020 | ···················· | ···················· | ····················· | ···················· | ···················· | ···················· |
| Z | 6027 hCoV-19/Wales/PHWC-49D21D/2020\|EPI_ISL_742751\|2020 | ···················· | ···················· | ····················· | ···················· | ···················· | ···················· |
| Z | 6028 hCoV-19/Wales/PHWC-49D22C/2020\|EPI_ISL_742752\|2020 | ···················· | ···················· | ····················· | ···················· | ···················· | ···················· |
| Z | 6029 hCoV-19/Wales/PHWC-49D24A/2020\|EPI_ISL_742754\|2020 | ···················· | ···················· | ····················· | ···················· | ···················· | ···················· |
| Z | 6030 hCoV-19/Wales/PHWC-49D259/2020\|EPI_ISL_742755\|2020 | ···················· | ···················· | ····················· | ···················· | ···················· | ···················· |
| Z | 6031 hCoV-19/Wales/PHWC-49D277/2020\|EPI_ISL_742757\|2020 | ···················· | ···················· | ····················· | ···················· | ···················· | ···················· |
| Z | 6032 hCoV-19/Wales/PHWC-49D286/2020\|EPI_ISL_742758\|2020 | ···················· | ···················· | ····················· | ···················· | ···················· | ···················· |
| Z | 6033 hCoV-19/Wales/PHWC-49D295/2020\|EPI_ISL_742759\|2020 | ···················· | ···················· | ····················· | ···················· | ···················· | ···················· |
| Z | 6034 hCoV-19/Wales/PHWC-49D2B3/2020\|EPI_ISL_742760\|2020 | ···················· | ···················· | ····················· | ···················· | ···················· | ···················· |
| Z | 6035 hCoV-19/Wales/PHWC-49D2C2/2020\|EPI_ISL_742761\|2020 | ···················· | ···················· | ····················· | ···················· | ···················· | ···················· |
| Z | 6036 hCoV-19/Wales/PHWC-49D2D1/2020\|EPI_ISL_742762\|2020 | ···················· | ···················· | ····················· | ···················· | ···················· | ···················· |
| Z | 6037 hCoV-19/Wales/PHWC-49D2E0/2020\|EPI_ISL_742763\|2020 | ···················· | ···················· | ····················· | ···················· | ···················· | ···················· |
| Z | 6038 hCoV-19/Wales/PHWC-49D2FF/2020\|EPI_ISL_742764\|2020 | ···················· | ···················· | ····················· | ···················· | ···················· | ···················· |
| Z | 6039 hCoV-19/Wales/PHWC-49D31A/2020\|EPI_ISL_742765\|2020 | ···················· | ···················· | ····················· | ···················· | ···················· | ···················· |
| Z | 6040 hCoV-19/Wales/PHWC-49D4EA/2020\|EPI_ISL_742792\|2020 | ···················· | ···················· | ····················· | ···················· | ···················· | ···················· |
| Z | 6041 hCoV-19/Wales/PHWC-49D4F9/2020\|EPI_ISL_742793\|2020 | ···················· | ···················· | ····················· | ···················· | ···················· | ···················· |
| Z | 6042 hCoV-19/Wales/PHWC-49D523/2020\|EPI_ISL_742795\|2020 | ···················· | ···················· | ····················· | ···················· | ···················· | ···················· |
| Z | 6043 hCoV-19/Wales/PHWC-49D532/2020\|EPI_ISL_742796\|2020 | ···················· | ···················· | ····················· | ···················· | ···················· | ···················· |
| Z | 6044 hCoV-19/Wales/PHWC-49D56F/2020\|EPI_ISL_742799\|2020 | ···················· | ···················· | ····················· | ···················· | ···················· | ···················· |
| Z | 6045 hCoV-19/USA/UT-UPHL-2012021070/2020\|EPI_ISL_746200 | ···················· | ·A·················· | ····················· | ···················· | ···················· | ···················· |
| Z | 6046 hCoV-19/USA/UT-UPHL-2012305257/2020\|EPI_ISL_746201 | ···················· | ···················· | ····················· | ···················· | ···················· | ···················· |
| Z | 6047 hCoV-19/USA/UT-UPHL-2012525389/2020\|EPI_ISL_746202 | ···················· | ···················· | ····················· | ···················· | ···················· | ···················· |
| Z | 6048 hCoV-19/USA/UT-UPHL-2012218264/2020\|EPI_ISL_746203 | ···················· | ···················· | ····················· | ···················· | ···················· | ···················· |
| Z | 6049 hCoV-19/USA/UT-UPHL-2012988344/2020\|EPI_ISL_746204 | ···················· | ···················· | ····················· | ···················· | ···················· | ···················· |
| Z | 6050 hCoV-19/USA/UT-UPHL-2012106449/2020\|EPI_ISL_746205 | ···················· | ···················· | ····················· | ···················· | ···················· | ···············A···· |
| Z | 6051 hCoV-19/USA/UT-UPHL-2012056833/2020\|EPI_ISL_746206 | ···················· | ···················· | ····················· | ···················· | ···················· | ···················· |
| Z | 6052 hCoV-19/USA/UT-UPHL-2012000168/2020\|EPI_ISL_746207 | ···················· | ·A·················· | ····················· | ···················· | ···················· | ···················· |
| Z | 6053 hCoV-19/USA/UT-UPHL-2012215761/2020\|EPI_ISL_746208 | ···················· | ···················· | ····················· | ···················· | ···················· | ···················· |
| Z | 6054 hCoV-19/USA/UT-UPHL-2012290335/2020\|EPI_ISL_746209 | ···················· | ···················· | ····················· | ···················· | ···················· | ···················· |
| Z | 6055 hCoV-19/USA/UT-UPHL-2012592285/2020\|EPI_ISL_746210 | ···················· | ·A·················· | ····················· | ···················· | ···················· | ···················· |
| Z | 6056 hCoV-19/USA/UT-UPHL-2012244887/2020\|EPI_ISL_746211 | ···················· | ···················· | ····················· | ···················· | ···················· | ···················· |
| Z | 6057 hCoV-19/USA/UT-UPHL-2012343317/2020\|EPI_ISL_746212 | ···················· | ···················· | ····················· | ···················· | ···················· | ···················· |
| Z | 6058 hCoV-19/USA/UT-UPHL-2012988875/2020\|EPI_ISL_746213 | ···················· | ···················· | ····················· | ···················· | ···················· | ···················· |
| Z | 6059 hCoV-19/USA/UT-UPHL-2012636660/2020\|EPI_ISL_746214 | ···················· | ···················· | ····················· | ···················· | ···················· | ···················· |
| Z | 6060 hCoV-19/USA/UT-UPHL-2012116345/2020\|EPI_ISL_746215 | ···················· | ···················· | ····················· | ···················· | ···················· | ···················· |
| Z | 6061 hCoV-19/USA/UT-UPHL-2012255039/2020\|EPI_ISL_746216 | ···················· | ···················· | ····················· | ···················· | ···················· | ···················· |
| Z | 6062 hCoV-19/USA/UT-UPHL-2012089430/2020\|EPI_ISL_746217 | ···················· | ···················· | ····················· | ···················· | ···················· | ···················· |
| Z | 6063 hCoV-19/USA/UT-UPHL-2012149731/2020\|EPI_ISL_746218 | ···················· | ···················· | ····················· | ···················· | ···················· | ···················· |
| Z | 6064 hCoV-19/USA/UT-UPHL-2012125874/2020\|EPI_ISL_746219 | ···················· | ···················· | ····················· | ···················· | ···················· | ···················· |
| Z | 6065 hCoV-19/USA/UT-UPHL-2012724092/2020\|EPI_ISL_746220 | ···················· | ···················· | ····················· | ···················· | ···················· | ···················· |
| Z | 6066 hCoV-19/USA/UT-UPHL-2012466667/2020\|EPI_ISL_746221 | ···················· | ·A·················· | ····················· | ···················· | ···················· | ···················· |
| Z | 6067 hCoV-19/USA/UT-UPHL-2012687578/2020\|EPI_ISL_746222 | ···················· | ···················· | ····················· | ···················· | ···················· | ···················· |
| Z | 6068 hCoV-19/USA/UT-UPHL-2012576849/2020\|EPI_ISL_746223 | ···················· | ···················· | ····················· | ···················· | ···················· | ···················· |
| Z | 6069 hCoV-19/USA/UT-UPHL-2012561708/2020\|EPI_ISL_746224 | ···················· | ···················· | ····················· | ···················· | ···················· | ···················· |
| Z | 6070 hCoV-19/USA/UT-UPHL-2012608096/2020\|EPI_ISL_746225 | ···················· | ·A·················· | ····················· | ···················· | ···················· | ···················· |
| Z | 6071 hCoV-19/USA/UT-UPHL-2012069730/2020\|EPI_ISL_746226 | ···················· | ···················· | ····················· | ···················· | ···················· | ···················· |
| Z | 6072 hCoV-19/USA/UT-UPHL-2012238828/2020\|EPI_ISL_746227 | ···················· | ···················· | ····················· | ···················· | ···················· | ···················· |
| Z | 6073 hCoV-19/USA/UT-UPHL-2012973122/2020\|EPI_ISL_746228 | ···················· | ···················· | ····················· | ···················· | ···················· | ···················· |
| Z | 6074 hCoV-19/USA/UT-UPHL-2012732813/2020\|EPI_ISL_746229 | ···················· | ···················· | ····················· | ···················· | ···················· | ···················· |
| Z | 6075 hCoV-19/USA/UT-UPHL-2012399168/2020\|EPI_ISL_746230 | ···················· | ···················· | ····················· | ···················· | ···················· | ···················· |
| Z | 6076 hCoV-19/USA/UT-UPHL-2012361388/2020\|EPI_ISL_746231 | ···················· | ·A·················· | ····················· | ···················· | ···················· | ···················· |
| Z | 6077 hCoV-19/USA/UT-UPHL-2012864954/2020\|EPI_ISL_746232 | ···················· | ···················· | ····················· | ···················· | ···················· | ···················· |
| Z | 6078 hCoV-19/USA/UT-UPHL-2012876879/2020\|EPI_ISL_746233 | ···················· | ···················· | ····················· | ···················· | ···················· | ···················· |
| Z | 6079 hCoV-19/USA/UT-UPHL-2012994965/2020\|EPI_ISL_746234 | ···················· | ···················· | ····················· | ···················· | ···················· | ···················· |
| Z | 6080 hCoV-19/USA/UT-UPHL-2012427069/2020\|EPI_ISL_746235 | ···················· | ·A·················· | ····················· | ···················· | ···················· | ···················· |
| Z | 6081 hCoV-19/USA/UT-UPHL-2012500123/2020\|EPI_ISL_746236 | ···················· | ···················· | ····················· | ···················· | ···················· | ···················· |
| Z | 6082 hCoV-19/USA/UT-UPHL-2012555879/2020\|EPI_ISL_746237 | ···················· | ···················· | ····················· | ···················· | ···················· | ···················· |
| Z | 6083 hCoV-19/USA/UT-UPHL-2012527409/2020\|EPI_ISL_746238 | ···················· | ···················· | ····················· | ···················· | ···················· | ···················· |
| Z | 6084 hCoV-19/USA/UT-UPHL-2012403664/2020\|EPI_ISL_746239 | ···················· | ···················· | ····················· | ···················· | ···················· | ···················· |
| Z | 6085 hCoV-19/USA/UT-UPHL-2012379360/2020\|EPI_ISL_746240 | ···················· | ···················· | ····················· | ···················· | ···················· | ···················· |
| Z | 6086 hCoV-19/USA/UT-UPHL-2012505195/2020\|EPI_ISL_746241 | ···················· | ···················· | ····················· | ···················· | ···················· | ···················· |
| Z | 6087 hCoV-19/USA/UT-UPHL-2012818790/2020\|EPI_ISL_746242 | ···················· | ···················· | ····················· | ···················· | ···················· | ···················· |
| Z | 6088 hCoV-19/USA/UT-UPHL-2012623393/2020\|EPI_ISL_746243 | ···················· | ···················· | ····················· | ···················· | ···················· | ···················· |
| Z | 6089 hCoV-19/USA/UT-UPHL-2012340933/2020\|EPI_ISL_746244 | ···················· | ···················· | ····················· | ···················· | ···················· | ···················· |
| Z | 6090 hCoV-19/USA/UT-UPHL-2012773673/2020\|EPI_ISL_746245 | ···················· | ···················· | ····················· | ···················· | ···················· | ···················· |
| Z | 6091 hCoV-19/USA/UT-UPHL-2012506772/2020\|EPI_ISL_746246 | ···················· | ···················· | ····················· | ···················· | ···················· | ···················· |
| Z | 6092 hCoV-19/USA/UT-UPHL-2012782938/2020\|EPI_ISL_746247 | ···················· | ···················· | ····················· | ···················· | ···················· | ···················· |
| Z | 6093 hCoV-19/USA/UT-UPHL-2012582863/2020\|EPI_ISL_746248 | ···················· | ···················· | ····················· | ···················· | ···················· | ···················· |
| Z | 6094 hCoV-19/USA/UT-UPHL-2012538969/2020\|EPI_ISL_746249 | ···················· | ···················· | ····················· | ···················· | ···················· | ···················· |
| Z | 6095 hCoV-19/USA/UT-UPHL-2012765469/2020\|EPI_ISL_746250 | ···················· | ···················· | ····················· | ···················· | ···················· | ···················· |
| Z | 6096 hCoV-19/USA/UT-UPHL-2012275795/2020\|EPI_ISL_746251 | ···················· | ···················· | ····················· | ···················· | ···················· | ···················· |
| Z | 6097 hCoV-19/USA/UT-UPHL-2012375866/2020\|EPI_ISL_746252 | ···················· | ···················· | ····················· | ···················· | ···················· | ···················· |
| Z | 6098 hCoV-19/USA/UT-UPHL-2012588723/2020\|EPI_ISL_746253 | ···················· | ···················· | ····················· | ···················· | ···················· | ···············A···· |
| Z | 6099 hCoV-19/USA/UT-UPHL-2012315956/2020\|EPI_ISL_746254 | ···················· | ···················· | ····················· | ···················· | ···················· | ···················· |

| | | | | | | | | |
|---|---|---|---|---|---|---|---|---|
| Z | 6100 | hCoV-19/USA/UT-UPHL-2012803704/2020\|EPI_ISL_746255 | ···················· | ·A·················· | ···················· | ···················· | ···················· | ···················· |
| Z | 6101 | hCoV-19/USA/UT-UPHL-2012394987/2020\|EPI_ISL_746256 | ···················· | ·A·················· | ···················· | ···················· | ···················· | ···················· |
| Z | 6102 | hCoV-19/USA/UT-UPHL-2012118383/2020\|EPI_ISL_746257 | ···················· | ···················· | ···················· | ···················· | ···················· | ···················· |
| Z | 6103 | hCoV-19/USA/UT-UPHL-2012278413/2020\|EPI_ISL_746258 | ···················· | ···················· | ···················· | ···················· | ···················· | ···················· |
| Z | 6104 | hCoV-19/USA/UT-UPHL-2012921757/2020\|EPI_ISL_746259 | ···················· | ···················· | ···················· | ···················· | ···················· | ···················· |
| Z | 6105 | hCoV-19/USA/UT-UPHL-2012280511/2020\|EPI_ISL_746260 | ···················· | ···················· | ···················· | ···················· | ···················· | ···················· |
| Z | 6106 | hCoV-19/USA/UT-UPHL-2012190007/2020\|EPI_ISL_746261 | ···················· | ···················· | ···················· | ···················· | ···················· | ···················· |
| Z | 6107 | hCoV-19/USA/UT-UPHL-2012926511/2020\|EPI_ISL_746262 | ···················· | ···················· | ···················· | ···················· | ···················· | ···················· |
| Z | 6108 | hCoV-19/USA/UT-UPHL-2012434776/2020\|EPI_ISL_746263 | ···················· | ·A·················· | ···················· | ···················· | ···················· | ···················· |
| Z | 6109 | hCoV-19/USA/UT-UPHL-2012956729/2020\|EPI_ISL_746264 | ···················· | ·A·················· | ···················· | ···················· | ···················· | ···················· |
| Z | 6110 | hCoV-19/USA/UT-UPHL-2012375605/2020\|EPI_ISL_746265 | ···················· | ···················· | ···················· | ···················· | ···················· | ···················· |
| Z | 6111 | hCoV-19/USA/UT-UPHL-2012983488/2020\|EPI_ISL_746266 | ···················· | ···················· | ···················· | ···················· | ···················· | ···················· |
| Z | 6112 | hCoV-19/USA/UT-UPHL-2012827102/2020\|EPI_ISL_746267 | ···················· | ···················· | ···················· | ···················· | ···················· | ···················· |
| Z | 6113 | hCoV-19/USA/UT-UPHL-2012100868/2020\|EPI_ISL_746268 | ···················· | ·A·················· | ···················· | ···················· | ···················· | ···················· |
| Z | 6114 | hCoV-19/USA/UT-UPHL-2012264814/2020\|EPI_ISL_746269 | ···················· | ···················· | ···················· | ···················· | ···················· | ···················· |
| Z | 6115 | hCoV-19/USA/UT-UPHL-2012682075/2020\|EPI_ISL_746270 | ···················· | ···················· | ···················· | ···················· | ···················· | ···················· |
| Z | 6116 | hCoV-19/USA/UT-UPHL-2012130640/2020\|EPI_ISL_746271 | ···················· | ···················· | ···················· | ···················· | ···················· | ···················· |
| Z | 6117 | hCoV-19/USA/UT-UPHL-2012218996/2020\|EPI_ISL_746272 | ···················· | ···················· | ···················· | ···················· | ···················· | ···················· |
| Z | 6118 | hCoV-19/USA/UT-UPHL-2012118876/2020\|EPI_ISL_746273 | ···················· | ···················· | ···················· | ···················· | ···················· | ···················· |
| Z | 6119 | hCoV-19/USA/UT-UPHL-2012359598/2020\|EPI_ISL_746274 | ···················· | ·A·················· | ···················· | ···················· | ···················· | ···················· |
| Z | 6120 | hCoV-19/USA/UT-UPHL-2012360496/2020\|EPI_ISL_746275 | ···················· | ···················· | ···················· | ···················· | ···················· | ···················· |
| Z | 6121 | hCoV-19/USA/UT-UPHL-2012852183/2020\|EPI_ISL_746276 | ···················· | ···················· | ···················· | ···················· | ···················· | ···················· |
| Z | 6122 | hCoV-19/USA/UT-UPHL-2012197253/2020\|EPI_ISL_746277 | ···················· | ···················· | ···················· | ···················· | ······T············· | ···················· |
| Z | 6123 | hCoV-19/USA/UT-UPHL-2012388417/2020\|EPI_ISL_746278 | ···················· | ·A·················· | ···················· | ···················· | ···················· | ···················· |
| Z | 6124 | hCoV-19/USA/UT-UPHL-2012204862/2020\|EPI_ISL_746279 | ···················· | ···················· | ···················· | ···················· | ···················· | ···················· |
| Z | 6125 | hCoV-19/USA/UT-UPHL-2012559004/2020\|EPI_ISL_746280 | ···················· | ·A·················· | ···················· | ···················· | ···················· | ···················· |
| Z | 6126 | hCoV-19/USA/UT-UPHL-2012625321/2020\|EPI_ISL_746281 | ···················· | ···················· | ···················· | ···················· | ···················· | ···················· |
| Z | 6127 | hCoV-19/USA/UT-UPHL-2012414451/2020\|EPI_ISL_746282 | ···················· | ···················· | ···················· | ···················· | ···················· | ···················· |
| Z | 6128 | hCoV-19/USA/UT-UPHL-2012036570/2020\|EPI_ISL_746283 | ···················· | ···················· | ···················· | ···················· | ···················· | ···················· |
| Z | 6129 | hCoV-19/USA/UT-UPHL-2012776444/2020\|EPI_ISL_746284 | ···················· | ···················· | ···················· | ···················· | ···················· | ···················· |
| Z | 6130 | hCoV-19/USA/UT-UPHL-2012590975/2020\|EPI_ISL_746285 | ···················· | ···················· | ···················· | ···················· | ···················· | ···················· |
| Z | 6131 | hCoV-19/USA/UT-UPHL-2012460400/2020\|EPI_ISL_746286 | ···················· | ···················· | ···················· | ···················· | ···················· | ···················· |
| Z | 6132 | hCoV-19/USA/UT-UPHL-2012739315/2020\|EPI_ISL_746287 | ···················· | ·A·················· | ···················· | ···················· | ···················· | ···················· |
| Z | 6133 | hCoV-19/USA/UT-UPHL-2012310170/2020\|EPI_ISL_746288 | ···················· | ···················· | ···················· | ···················· | ···················· | ···················· |
| Z | 6134 | hCoV-19/USA/UT-UPHL-2012703634/2020\|EPI_ISL_746289 | ···················· | ···················· | ···················· | ···················· | ···················· | ···················· |
| Z | 6135 | hCoV-19/USA/UT-UPHL-2012148746/2020\|EPI_ISL_746290 | ···················· | ···················· | ···················· | ···················· | ···················· | ···············A···· |
| Z | 6136 | hCoV-19/USA/UT-UPHL-2012328423/2020\|EPI_ISL_746291 | ···················· | ···················· | ···················· | ···················· | ···················· | ···················· |
| Z | 6137 | hCoV-19/USA/UT-UPHL-2012550362/2020\|EPI_ISL_746292 | ···················· | ·A·················· | ···················· | ···················· | ···················· | ···················· |
| Z | 6138 | hCoV-19/USA/UT-UPHL-2012133022/2020\|EPI_ISL_746293 | ···················· | ···················· | ···················· | ···················· | ···················· | ···················· |
| Z | 6139 | hCoV-19/USA/UT-UPHL-2012275073/2020\|EPI_ISL_746294 | ···················· | ···················· | ···················· | ···················· | ···················· | ···················· |
| Z | 6140 | hCoV-19/USA/UT-UPHL-2012424387/2020\|EPI_ISL_746295 | ···················· | ···················· | ···················· | ···················· | ···················· | ···················· |
| Z | 6141 | hCoV-19/USA/UT-UPHL-2012883694/2020\|EPI_ISL_746296 | ···················· | ···················· | ···················· | ···················· | ···················· | ···················· |
| Z | 6142 | hCoV-19/USA/UT-UPHL-2012190312/2020\|EPI_ISL_746297 | ···················· | ···················· | ···················· | ···················· | ···················· | ···················· |
| Z | 6143 | hCoV-19/USA/UT-UPHL-2012351195/2020\|EPI_ISL_746298 | ···················· | ···················· | ···················· | ···················· | ···················· | ···················· |
| Z | 6144 | hCoV-19/USA/UT-UPHL-2012721054/2020\|EPI_ISL_746299 | ···················· | ···················· | ···················· | ···················· | ···················· | ···················· |
| Z | 6145 | hCoV-19/USA/CA-CZB-13966/2020\|EPI_ISL_739120\|2020- | ···················· | ···················· | ···················· | ···················· | ···················· | ···················· |
| Z | 6146 | hCoV-19/USA/CA-CZB-15179/2020\|EPI_ISL_739121\|2020- | ···················· | ·A·················· | ···················· | ···················· | ···················· | ···················· |
| Z | 6148 | hCoV-19/USA/CA-CZB-14832/2020\|EPI_ISL_739123\|2020- | ···················· | ···················· | ···················· | ···················· | ···················· | ···················· |
| Z | 6149 | hCoV-19/USA/CA-CZB-14403/2020\|EPI_ISL_739124\|2020- | ···················· | ···················· | ···················· | ···················· | ···················· | ···················· |
| Z | 6150 | hCoV-19/USA/CA-CZB-14895/2020\|EPI_ISL_739125\|2020- | ···················· | ···················· | ···················· | ···················· | ···················· | ···················· |
| Z | 6151 | hCoV-19/USA/CA-CZB-14854/2020\|EPI_ISL_739126\|2020- | ···················· | ···················· | ···················· | ···················· | ···················· | ···················· |
| Z | 6152 | hCoV-19/USA/CA-CZB-13734/2020\|EPI_ISL_739127\|2020- | ···················· | ·A·················· | ···················· | ···················· | ···················· | ···················· |
| Z | 6153 | hCoV-19/USA/CA-CZB-15144/2020\|EPI_ISL_739128\|2020- | ···················· | ·A·················· | ···················· | ···················· | ···················· | ···················· |
| Z | 6154 | hCoV-19/USA/CA-CZB-15098/2020\|EPI_ISL_739129\|2020- | ···················· | ···················· | ···················· | ···················· | ···················· | ···················· |
| Z | 6155 | hCoV-19/USA/CA-CZB-14545/2020\|EPI_ISL_739130\|2020- | ···················· | ···················· | ···················· | ···················· | ···················· | ···················· |
| Z | 6156 | hCoV-19/USA/CA-CZB-14865/2020\|EPI_ISL_739131\|2020- | ···················· | ···················· | ···················· | ···················· | ···················· | ···················· |
| Z | 6157 | hCoV-19/USA/CA-CZB-14946/2020\|EPI_ISL_739132\|2020- | ···················· | ···················· | ···················· | ···················· | ···················· | ···················· |
| Z | 6158 | hCoV-19/USA/CA-CZB-14153/2020\|EPI_ISL_739133\|2020- | ···················· | ···················· | ···················· | ···················· | ···················· | ···················· |
| Z | 6159 | hCoV-19/USA/CA-CZB-14347/2020\|EPI_ISL_739134\|2020- | ···················· | ·A·················· | ···················· | ···················· | ···················· | ···················· |
| Z | 6160 | hCoV-19/USA/CA-CZB-13973/2020\|EPI_ISL_739135\|2020- | ···················· | ···················· | ···················· | ···················· | ···················· | ···················· |
| Z | 6161 | hCoV-19/USA/CA-CZB-13824/2020\|EPI_ISL_739136\|2020- | ···················· | ···················· | ···················· | ···················· | ···················· | ···················· |
| Z | 6163 | hCoV-19/USA/CA-CZB-15158/2020\|EPI_ISL_739138\|2020- | ···················· | ·A·················· | ···················· | ···················· | ···················· | ···················· |
| Z | 6164 | hCoV-19/USA/CA-CZB-14631/2020\|EPI_ISL_739139\|2020- | ···················· | ···················· | ···················· | ···················· | ···················· | ···················· |
| Z | 6165 | hCoV-19/USA/CA-CZB-14918/2020\|EPI_ISL_739142\|2020- | ···················· | ···················· | ···················· | ···················· | ···················· | ···················· |
| Z | 6166 | hCoV-19/USA/CA-CZB-13727/2020\|EPI_ISL_739143\|2020- | ···················· | ···················· | ···················· | ···················· | ···················· | ···················· |
| Z | 6167 | hCoV-19/USA/CA-CZB-14057/2020\|EPI_ISL_739144\|2020- | ···················· | ···················· | ···················· | ···················· | ···················· | ···················· |
| Z | 6169 | hCoV-19/USA/CA-CZB-15282/2020\|EPI_ISL_739146\|2020- | ···················· | ···················· | ···················· | ···················· | ···················· | ···················· |
| Z | 6170 | hCoV-19/USA/CA-CZB-14977/2020\|EPI_ISL_739147\|2020- | ···················· | ···················· | ···················· | ···················· | ···················· | ···················· |
| Z | 6171 | hCoV-19/USA/CA-CZB-14752/2020\|EPI_ISL_739148\|2020- | ···················· | ···················· | ···················· | ···················· | ···················· | ···················· |
| Z | 6172 | hCoV-19/USA/CA-CZB-13967/2020\|EPI_ISL_739149\|2020- | ···················· | ···················· | ···················· | ···················· | ···················· | ···················· |
| Z | 6175 | hCoV-19/USA/CA-CZB-14694/2020\|EPI_ISL_739162\|2020- | ···················· | ···················· | ···················· | ···················· | ···················· | ···················· |
| Z | 6176 | hCoV-19/USA/CA-CZB-13885/2020\|EPI_ISL_739163\|2020- | ···················· | ·A·················· | ···················· | ···················· | ···················· | ···················· |
| Z | 6177 | hCoV-19/USA/CA-CZB-14088/2020\|EPI_ISL_739164\|2020- | ···················· | ···················· | ···················· | ···················· | ···················· | ···················· |
| Z | 6178 | hCoV-19/USA/CA-CZB-14763/2020\|EPI_ISL_739165\|2020- | ···················· | ···················· | ···················· | ···················· | ···················· | ···················· |
| Z | 6179 | hCoV-19/USA/CA-CZB-15960/2020\|EPI_ISL_739166\|2020- | ···················· | ·A·················· | ···················· | ···················· | ···················· | ···················· |
| Z | 6180 | hCoV-19/USA/CA-CZB-14531/2020\|EPI_ISL_739167\|2020- | ···················· | ·A·················· | ···················· | ···················· | ···················· | ···················· |
| Z | 6181 | hCoV-19/USA/CA-CZB-14532/2020\|EPI_ISL_739168\|2020- | ···················· | ·A·················· | ···················· | ···················· | ···················· | ···················· |
| Z | 6182 | hCoV-19/USA/CA-CZB-11270/2020\|EPI_ISL_739169\|2020- | ···················· | ···················· | ···················· | ···················· | ···················· | ···················· |
| Z | 6183 | hCoV-19/USA/CA-CZB-14816/2020\|EPI_ISL_739170\|2020- | ···················· | ···················· | ···················· | ···················· | ···················· | ···················· |
| Z | 6184 | hCoV-19/USA/CA-CZB-14110/2020\|EPI_ISL_739171\|2020- | ···················· | ···················· | ···················· | ···················· | ···················· | ···················· |

| | | | | | | | | |
|---|---|---|---|---|---|---|---|---|
| Z | 6185 | hCoV-19/USA/CA-CZB-14165/2020\|EPI_ISL_739172\|2020- | ···················· | ···················· | ···················· | ···················· | ···················· | ···················· |
| Z | 6186 | hCoV-19/USA/CA-CZB-13742/2020\|EPI_ISL_739173\|2020- | ···················· | ·A·················· | ···················· | ···················· | ···················· | ···················· |
| Z | 6188 | hCoV-19/USA/CA-CZB-14226/2020\|EPI_ISL_739175\|2020- | ···················· | ···················· | ···················· | ···················· | ···················· | ···················· |
| Z | 6189 | hCoV-19/USA/CA-CZB-15462/2020\|EPI_ISL_739176\|2020- | ···················· | ···················· | ··T················· | ···················· | ···················· | ···················· |
| Z | 6190 | hCoV-19/USA/CA-CZB-14741/2020\|EPI_ISL_739177\|2020- | ···················· | ·A·················· | ···········T········ | ···················· | ···················· | ···················· |
| Z | 6191 | hCoV-19/USA/CA-CZB-14144/2020\|EPI_ISL_739178\|2020- | ···················· | ···················· | ···················· | ···················· | ···················· | ···················· |
| Z | 6192 | hCoV-19/USA/CA-CZB-14306/2020\|EPI_ISL_739179\|2020- | ···················· | ·A·················· | ···················· | ···················· | ···················· | ···················· |
| Z | 6193 | hCoV-19/USA/CA-CZB-15190/2020\|EPI_ISL_739180\|2020- | ···················· | ···················· | ···················· | ···················· | ···················· | ···················· |
| Z | 6194 | hCoV-19/USA/CA-CZB-11272/2020\|EPI_ISL_739181\|2020- | ···················· | ···················· | ···················· | ···················· | ···················· | ···················· |
| Z | 6195 | hCoV-19/USA/CA-CZB-15446/2020\|EPI_ISL_739182\|2020- | ···················· | ···················· | ···················· | ···················· | ···················· | ···················· |
| Z | 6196 | hCoV-19/USA/CA-CZB-14842/2020\|EPI_ISL_739183\|2020- | ···················· | ···················· | ···················· | ···················· | ···················· | ···················· |
| Z | 6198 | hCoV-19/USA/CA-CZB-13729/2020\|EPI_ISL_739185\|2020- | ···················· | ···················· | ···················· | ···················· | ···················· | ···················· |
| Z | 6199 | hCoV-19/USA/CA-CZB-15968/2020\|EPI_ISL_739186\|2020- | ···················· | ·A·················· | ···················· | ···················· | ···················· | ···················· |
| Z | 6200 | hCoV-19/USA/CA-CZB-15115/2020\|EPI_ISL_739187\|2020- | ···················· | ···················· | ···················· | ···················· | ···················· | ···················· |
| Z | 6201 | hCoV-19/USA/CA-CZB-13668/2020\|EPI_ISL_739188\|2020- | ···················· | ·A·················· | ···················· | ···················· | ···················· | ···················· |
| Z | 6202 | hCoV-19/USA/CA-CZB-14411/2020\|EPI_ISL_739189\|2020- | ···················· | ·A·················· | ···················· | ···················· | ···················· | ···················· |
| Z | 6204 | hCoV-19/USA/CA-CZB-14910/2020\|EPI_ISL_739191\|2020- | ···················· | ···················· | ···················· | ···················· | ···················· | ···················· |
| Z | 6205 | hCoV-19/USA/CA-CZB-14131/2020\|EPI_ISL_739192\|2020- | ···················· | ···················· | ···················· | ···················· | ···················· | ···················· |
| Z | 6206 | hCoV-19/USA/CA-CZB-14713/2020\|EPI_ISL_739193\|2020- | ········T··········· | ···················· | ···················· | ···················· | ···················· | ···················· |
| Z | 6207 | hCoV-19/USA/CA-CZB-14788/2020\|EPI_ISL_739194\|2020- | ···················· | ···················· | ···················· | ···················· | ···················· | ···················· |
| Z | 6208 | hCoV-19/USA/CA-CZB-14577/2020\|EPI_ISL_739195\|2020- | ···················· | ···················· | ···················· | ···················· | ···················· | ···················· |
| Z | 6209 | hCoV-19/USA/CA-CZB-14133/2020\|EPI_ISL_739196\|2020- | ···················· | ···················· | ···················· | ···················· | ···················· | ···················· |
| Z | 6210 | hCoV-19/USA/CA-CZB-14154/2020\|EPI_ISL_739197\|2020- | ···················· | ·A·················· | ···················· | ···················· | ···················· | ···················· |
| Z | 6211 | hCoV-19/USA/CA-CZB-14862/2020\|EPI_ISL_739198\|2020- | ···················· | ···················· | ···················· | ···················· | ···················· | ···················· |
| Z | 6212 | hCoV-19/USA/CA-CZB-14230/2020\|EPI_ISL_739199\|2020- | ···················· | ···················· | ···················· | ···················· | ·NN················· | ···················· |
| Z | 6213 | hCoV-19/Luxembourg/LNS2353921/2020\|EPI_ISL_740104\| | ···················· | ···················· | ···················· | ···················· | ···················· | ···················· |
| Z | 6214 | hCoV-19/Luxembourg/LNS2356539/2020\|EPI_ISL_740106\| | ···················· | ···················· | ···················· | ···················· | ···················· | ···················· |
| Z | 6215 | hCoV-19/Luxembourg/LNS2360705/2020\|EPI_ISL_740107\| | ···················· | ···················· | ···················· | ···················· | ···················· | ···················· |
| Z | 6216 | hCoV-19/Luxembourg/LNS2363167/2020\|EPI_ISL_740108\| | ···················· | ···················· | ···················· | ···················· | ···················· | ···················· |
| Z | 6217 | hCoV-19/Luxembourg/LNS2364456/2020\|EPI_ISL_740109\| | ···········T········ | ···················· | ···················· | ···················· | ···················· | ···················· |
| Z | 6218 | hCoV-19/Luxembourg/LNS2370192/2020\|EPI_ISL_740110\| | ···················· | ···················· | ···················· | ···················· | ···················· | ···················· |
| Z | 6219 | hCoV-19/Luxembourg/LNS2388068/2020\|EPI_ISL_740111\| | ···················· | ···················· | ···················· | ···················· | ···················· | ···················· |
| Z | 6220 | hCoV-19/Luxembourg/LNS2390229/2020\|EPI_ISL_740112\| | ···················· | ···················· | ···················· | ···················· | ···················· | ···················· |
| Z | 6221 | hCoV-19/Luxembourg/LNS2391079/2020\|EPI_ISL_740113\| | ···················· | ···················· | ···················· | ···················· | ···················· | ···················· |
| Z | 6222 | hCoV-19/Luxembourg/LNS2399095/2020\|EPI_ISL_740114\| | ···················· | ···················· | ···················· | ···················· | ···················· | ···················· |
| Z | 6223 | hCoV-19/Luxembourg/LNS2402544/2020\|EPI_ISL_740115\| | ···················· | ···················· | ···················· | ···················· | ···················· | ···················· |
| Z | 6224 | hCoV-19/Luxembourg/LNS2402896/2020\|EPI_ISL_740116\| | ···················· | ···················· | ···················· | ···················· | ······T············· | ···················· |
| Z | 6225 | hCoV-19/Luxembourg/LNS2404691/2020\|EPI_ISL_740117\| | ···················· | ···················· | ···················· | ···················· | ···················· | ···················· |
| Z | 6226 | hCoV-19/Luxembourg/LNS2407829/2020\|EPI_ISL_740118\| | ···················· | ···················· | ···················· | ···················· | ···················· | ···················· |
| Z | 6227 | hCoV-19/Luxembourg/LNS2409206/2020\|EPI_ISL_740119\| | ···················· | ···················· | ···················· | ···················· | ···················· | ···················· |
| Z | 6228 | hCoV-19/Luxembourg/LNS2424302/2020\|EPI_ISL_740120\| | ···················· | ···················· | ···················· | ···················· | ···················· | ···················· |
| Z | 6229 | hCoV-19/Luxembourg/LNS2425583/2020\|EPI_ISL_740121\| | ···················· | ···················· | ···················· | ···················· | ···················· | ···················· |
| Z | 6230 | hCoV-19/Luxembourg/LNS2433992/2020\|EPI_ISL_740122\| | ···················· | ···················· | ···················· | ···················· | ···················· | ···················· |
| Z | 6231 | hCoV-19/Luxembourg/LNS2435517/2020\|EPI_ISL_740123\| | ···················· | ···················· | ···················· | ···················· | ···················· | ···················· |
| Z | 6232 | hCoV-19/Luxembourg/LNS2444455/2020\|EPI_ISL_740124\| | ···················· | ···················· | ···················· | ···················· | ···················· | ···················· |
| Z | 6233 | hCoV-19/Luxembourg/LNS2447440/2020\|EPI_ISL_740126\| | ···················· | ···················· | ···················· | ···················· | ···················· | ···················· |
| Z | 6234 | hCoV-19/Luxembourg/LNS2485743/2020\|EPI_ISL_740128\| | ···················· | ····A··············· | ···················· | ···················· | ···················· | ···················· |
| Z | 6235 | hCoV-19/Luxembourg/LNS2520681/2020\|EPI_ISL_740134\| | ···················· | ···················· | ···················· | ···················· | ···················· | ···················· |
| Z | 6236 | hCoV-19/Luxembourg/LNS2523106/2020\|EPI_ISL_740135\| | ···················· | ···················· | ···················· | ···················· | ···················· | ···················· |
| Z | 6237 | hCoV-19/Luxembourg/LNS2529586/2020\|EPI_ISL_740136\| | ···················· | ···················· | ···················· | ···················· | ···················· | ···················· |
| Z | 6238 | hCoV-19/Luxembourg/LNS2589258/2020\|EPI_ISL_740148\| | ···················· | ···················· | ···················· | ···················· | ······T············· | ···················· |
| Z | 6239 | hCoV-19/Luxembourg/LNS2593581/2020\|EPI_ISL_740149\| | ···················· | ···················· | ···················· | ···················· | ···················· | ···················· |
| Z | 6240 | hCoV-19/Luxembourg/LNS2594950/2020\|EPI_ISL_740151\| | ···················· | ····A··············· | ···················· | ···················· | ···················· | ···················· |
| Z | 6241 | hCoV-19/Luxembourg/LNS2596234/2020\|EPI_ISL_740152\| | ···················· | ···················· | ···················· | ···················· | ···················· | ···················· |
| Z | 6242 | hCoV-19/Luxembourg/LNS2597209/2020\|EPI_ISL_740153\| | ···················· | ···················· | ···················· | ···················· | ···················· | ···················· |
| Z | 6243 | hCoV-19/Luxembourg/LNS2599024/2020\|EPI_ISL_740154\| | ···················· | ···················· | ···················· | ···················· | ···················· | ···················· |
| Z | 6244 | hCoV-19/Luxembourg/LNS2599728/2020\|EPI_ISL_740155\| | ···················· | ···················· | ···················· | ···················· | ···················· | ···················· |
| Z | 6245 | hCoV-19/Luxembourg/LNS2601498/2020\|EPI_ISL_740156\| | ···················· | ···················· | ···················· | ···················· | ···················· | ···················· |
| Z | 6246 | hCoV-19/Luxembourg/LNS2611167/2020\|EPI_ISL_740158\| | ···················· | ·A·················· | ···················· | ···················· | ···················· | ···················· |
| Z | 6247 | hCoV-19/Luxembourg/LNS2618320/2020\|EPI_ISL_740159\| | ···················· | ···················· | ···················· | ···················· | ···················· | ···················· |
| Z | 6248 | hCoV-19/Luxembourg/LNS2632689/2020\|EPI_ISL_740160\| | ···················· | ···················· | ···················· | ···················· | ···················· | ···················· |
| Z | 6249 | hCoV-19/Luxembourg/LNS2642998/2020\|EPI_ISL_740161\| | ···················· | ···················· | ···················· | ···················· | ···················· | ···················· |
| Z | 6250 | hCoV-19/Luxembourg/LNS2644869/2020\|EPI_ISL_740162\| | ···················· | ···················· | ···················· | ···················· | ···················· | ···················· |
| Z | 6251 | hCoV-19/Luxembourg/LNS2653287/2020\|EPI_ISL_740163\| | ···················· | ···················· | ···················· | ···················· | ···················· | ···················· |
| Z | 6252 | hCoV-19/Luxembourg/LNS2669816/2020\|EPI_ISL_740164\| | ···················· | ···················· | ···················· | ···················· | ···················· | ···················· |
| Z | 6253 | hCoV-19/Luxembourg/LNS2676018/2020\|EPI_ISL_740165\| | ···················· | ···················· | ···················· | ···················· | ···················· | ···················· |
| Z | 6254 | hCoV-19/Luxembourg/LNS2677404/2020\|EPI_ISL_740166\| | ···················· | ···················· | ···················· | ···················· | ···················· | ···················· |
| Z | 6255 | hCoV-19/Luxembourg/LNS2680625/2020\|EPI_ISL_740167\| | ···················· | ···················· | ···················· | ···················· | ···················· | ···················· |
| Z | 6256 | hCoV-19/Luxembourg/LNS2687447/2020\|EPI_ISL_740168\| | ···················· | ···················· | ···················· | ···················· | ···················· | ···················· |
| Z | 6257 | hCoV-19/Luxembourg/LNS2689418/2020\|EPI_ISL_740169\| | ···················· | ···················· | ···················· | ···················· | ···················· | ···················· |
| Z | 6258 | hCoV-19/Luxembourg/LNS2691541/2020\|EPI_ISL_740170\| | ···················· | ···················· | ···················· | ···················· | ···················· | ···················· |
| Z | 6259 | hCoV-19/Luxembourg/LNS2696567/2020\|EPI_ISL_740171\| | ···················· | ···················· | ···················· | ···················· | ···················· | ···················· |
| Z | 6260 | hCoV-19/Luxembourg/LNS2699765/2020\|EPI_ISL_740172\| | ···················· | ···················· | ···················· | ···················· | ···················· | ···················· |
| Z | 6261 | hCoV-19/Luxembourg/LNS2702537/2020\|EPI_ISL_740173\| | ···················· | ···················· | ···················· | ···················· | ···················· | ···················· |
| Z | 6262 | hCoV-19/Luxembourg/LNS2705456/2020\|EPI_ISL_740174\| | ···················· | ···················· | ···················· | ···················· | ···················· | ···················· |
| Z | 6263 | hCoV-19/Luxembourg/LNS2705564/2020\|EPI_ISL_740175\| | ···················· | ···················· | ···················· | ···················· | ···················· | ···················· |
| Z | 6264 | hCoV-19/Luxembourg/LNS2710395/2020\|EPI_ISL_740177\| | ···················· | ···················· | ···········T········ | ···················· | ···················· | ···················· |
| Z | 6265 | hCoV-19/Luxembourg/LNS2721373/2020\|EPI_ISL_740178\| | ···················· | ···················· | ···················· | ···················· | ···················· | ···················· |
| Z | 6266 | hCoV-19/Luxembourg/LNS2730340/2020\|EPI_ISL_740179\| | ···················· | ···················· | ···················· | ···················· | ···················· | ···················· |
| Z | 6267 | hCoV-19/Luxembourg/LNS2730488/2020\|EPI_ISL_740180\| | ···················· | ···················· | ···················· | ···················· | ···················· | ···················· |

| | | | | | | | | |
|---|---|---|---|---|---|---|---|---|
| Z | 6268 hCoV-19/Luxembourg/LNS2740500/2020\|EPI_ISL_740181\| | ···················· | ···················· | ···················· | ···················· | ···················· | ···················· | |
| Z | 6269 hCoV-19/Luxembourg/LNS2749430/2020\|EPI_ISL_740183\| | ···················· | ···················· | ···················· | ···················· | ···················· | ···················· | |
| Z | 6270 hCoV-19/Luxembourg/LNS2751088/2020\|EPI_ISL_740184\| | ···················· | ···················· | ···················· | ···················· | ······T············· | ···················· | |
| Z | 6271 hCoV-19/Luxembourg/LNS2755277/2020\|EPI_ISL_740185\| | ···················· | ···················· | ···················· | ···················· | ···················· | ···················· | |
| Z | 6272 hCoV-19/Luxembourg/LNS2761686/2020\|EPI_ISL_740186\| | ···················· | ···················· | ···················· | ···················· | ···················· | ···················· | |
| Z | 6273 hCoV-19/Luxembourg/LNS2765469/2020\|EPI_ISL_740187\| | ···················· | ···················· | ···················· | ···················· | ···················· | ···················· | |
| Z | 6274 hCoV-19/Luxembourg/LNS2772835/2020\|EPI_ISL_740188\| | ···················· | ···················· | ···················· | ···················· | ···················· | ···················· | |
| Z | 6275 hCoV-19/Luxembourg/LNS2774762/2020\|EPI_ISL_740189\| | ···················· | ···················· | ···················· | ···················· | ···················· | ···················· | |
| Z | 6276 hCoV-19/Luxembourg/LNS2776151/2020\|EPI_ISL_740190\| | ···················· | ···················· | ···················· | ···················· | ···················· | ···················· | |
| Z | 6277 hCoV-19/Luxembourg/LNS2776427/2020\|EPI_ISL_740191\| | ···················· | ···················· | ···················· | ···················· | ···················· | ···················· | |
| Z | 6278 hCoV-19/Luxembourg/LNS2781126/2020\|EPI_ISL_740192\| | ···················· | ···················· | ···················· | ···················· | ···················· | ···················· | |
| Z | 6279 hCoV-19/Luxembourg/LNS2783388/2020\|EPI_ISL_740193\| | ···················· | ···················· | ···················· | ···················· | ······T············· | ···················· | |
| Z | 6280 hCoV-19/Luxembourg/LNS2791698/2020\|EPI_ISL_740194\| | ···················· | G··················· | ···················· | ···················· | ···················· | ···················· | |
| Z | 6281 hCoV-19/Luxembourg/LNS2794838/2020\|EPI_ISL_740195\| | ···················· | ···················· | ···················· | ···················· | ···················· | ···················· | |
| Z | 6282 hCoV-19/Luxembourg/LNS2795371/2020\|EPI_ISL_740196\| | ···················· | ···················· | ···················· | ···················· | ···················· | ···················· | |
| Z | 6283 hCoV-19/Luxembourg/LNS2797754/2020\|EPI_ISL_740197\| | ···················· | ···················· | ···················· | ···················· | ···················· | ···················· | |
| Z | 6284 hCoV-19/Luxembourg/LNS2803003/2020\|EPI_ISL_740198\| | ···················· | ···················· | ···················· | ···················· | ···················· | ···················· | |
| Z | 6285 hCoV-19/Luxembourg/LNS2813963/2020\|EPI_ISL_740199\| | ···················· | ···················· | ···················· | ···················· | ···················· | ···················· | |
| Z | 6286 hCoV-19/England/NORT-1B797F0/2020\|EPI_ISL_741436\|2 | ···················· | ···················· | ···················· | ···················· | ···················· | ···················· | |
| Z | 6287 hCoV-19/England/NORT-1B79839/2020\|EPI_ISL_741438\|2 | ···················· | ···················· | ···················· | ···················· | ···················· | ···················· | |
| Z | 6288 hCoV-19/England/NORT-1B79909/2020\|EPI_ISL_741440\|2 | ···················· | ···················· | ···················· | ···················· | ···················· | ···················· | |
| Z | 6289 hCoV-19/England/NORT-1B799CD/2020\|EPI_ISL_741441\|2 | ···················· | ···················· | ···················· | ···················· | ···················· | ···················· | |
| Z | 6290 hCoV-19/England/NORT-1B799FA/2020\|EPI_ISL_741442\|2 | ···················· | ···················· | ···················· | ···················· | ···················· | ···················· | |
| Z | 6291 hCoV-19/England/NORT-1B79A06/2020\|EPI_ISL_741443\|2 | ···················· | ···················· | ···················· | ···················· | ···················· | ···················· | |
| Z | 6292 hCoV-19/England/NORT-1B79A24/2020\|EPI_ISL_741444\|2 | ···················· | ···················· | ···················· | ···················· | ···················· | ···················· | |
| Z | 6293 hCoV-19/England/NORT-1B79ABB/2020\|EPI_ISL_741445\|2 | ···················· | ···················· | ···················· | ···················· | ···················· | ···················· | |
| Z | 6294 hCoV-19/England/NORT-1B79AE8/2020\|EPI_ISL_741447\|2 | ···················· | ···················· | ···················· | ···················· | ···················· | ···················· | |
| Z | 6295 hCoV-19/England/NORT-1B79AF7/2020\|EPI_ISL_741448\|2 | ···················· | ···················· | ···················· | ················T··· | ······T············· | ···················· | |
| Z | 6296 hCoV-19/England/NORT-1B79B5E/2020\|EPI_ISL_741451\|2 | ···················· | ···················· | ···················· | ···················· | ···················· | ···················· | |
| Z | 6297 hCoV-19/England/NORT-1B79B6D/2020\|EPI_ISL_741452\|2 | ···················· | ···················· | ···················· | ···················· | ···················· | ···················· | |
| Z | 6298 hCoV-19/England/NORW-21F989/2020\|EPI_ISL_741456\|20 | ···················· | ···················· | ···················· | ···················· | ···················· | ···················· | |
| Z | 6299 hCoV-19/England/NORW-21F998/2020\|EPI_ISL_741457\|20 | ···················· | ···················· | ···················· | ···················· | ···················· | ···················· | |
| Z | 6300 hCoV-19/England/NORW-21FA0E/2020\|EPI_ISL_741458\|20 | ···················· | ·A·················· | ···················· | ···················· | ···················· | ··A················· | |
| Z | 6301 hCoV-19/England/NORW-21FA3B/2020\|EPI_ISL_741459\|20 | ···················· | ···················· | ···················· | ···················· | ···················· | ···················· | |
| Z | 6302 hCoV-19/England/NORW-21FBCF/2020\|EPI_ISL_741466\|20 | ···················· | ·A·················· | ···················· | ···················· | ···················· | ··A················· | |
| Z | 6303 hCoV-19/England/NORW-21FBED/2020\|EPI_ISL_741467\|20 | ···················· | ···················· | ···················· | ···················· | ···················· | ···················· | |
| Z | 6304 hCoV-19/England/NORW-21FD8D/2020\|EPI_ISL_741479\|20 | ···················· | ···················· | ···················· | ···················· | ···················· | ···················· | |
| Z | 6305 hCoV-19/England/NORW-21FED5/2020\|EPI_ISL_741484\|20 | ···················· | ···················· | ···················· | ···················· | ···················· | ···················· | |
| Z | 6306 hCoV-19/England/NORW-21FF0F/2020\|EPI_ISL_741485\|20 | ···················· | ···················· | ···················· | ···················· | ···················· | ···················· | |
| Z | 6307 hCoV-19/England/NORW-220B25/2020\|EPI_ISL_741489\|20 | ···················· | ···················· | ···················· | ···················· | ···················· | ···················· | |
| Z | 6308 hCoV-19/England/NORW-220B8F/2020\|EPI_ISL_741490\|20 | ···················· | ·A·················· | ···················· | ···················· | ···················· | ··A················· | |
| Z | 6309 hCoV-19/England/NORW-220BE9/2020\|EPI_ISL_741491\|20 | ···················· | ···················· | ···················· | ···················· | ···················· | ···················· | |
| Z | 6310 hCoV-19/England/NORW-220C22/2020\|EPI_ISL_741493\|20 | ···················· | ···················· | ···················· | ···················· | ···················· | ···················· | |
| Z | 6311 hCoV-19/England/NORW-2223D0/2020\|EPI_ISL_741494\|20 | ···················· | ···················· | ···················· | ···················· | ···················· | ···················· | |
| Z | 6312 hCoV-19/England/NORW-2224BF/2020\|EPI_ISL_741496\|20 | ···················· | ···················· | ···················· | ···················· | ···················· | ···················· | |
| Z | 6313 hCoV-19/England/NORW-2224CE/2020\|EPI_ISL_741497\|20 | ···················· | ···················· | ···················· | ···················· | ···················· | ···················· | |
| Z | 6314 hCoV-19/England/NORW-222507/2020\|EPI_ISL_741498\|20 | ···················· | ···················· | ···················· | ···················· | ···················· | ···················· | |
| Z | 6315 hCoV-19/England/NORW-222516/2020\|EPI_ISL_741499\|20 | ···················· | ·A·················· | ···················· | ···················· | ···················· | ··A················· | |
| Z | 6316 hCoV-19/Wales/PHWC-49D338/2020\|EPI_ISL_742766\|2020 | ···················· | ···················· | ···················· | ···················· | ···················· | ···················· | |
| Z | 6317 hCoV-19/Wales/PHWC-49D365/2020\|EPI_ISL_742769\|2020 | ···················· | ···················· | ···················· | ···················· | ···················· | ···················· | |
| Z | 6318 hCoV-19/Wales/PHWC-49D374/2020\|EPI_ISL_742770\|2020 | ···················· | ···················· | ···················· | ···················· | ···················· | ···················· | |
| Z | 6319 hCoV-19/Wales/PHWC-49D383/2020\|EPI_ISL_742771\|2020 | ···················· | ·········A·········· | ···················· | ···················· | ···················· | ···················· | |
| Z | 6320 hCoV-19/Wales/PHWC-49D392/2020\|EPI_ISL_742772\|2020 | ···················· | ···················· | ···················· | ···················· | ···················· | ···················· | |
| Z | 6321 hCoV-19/Wales/PHWC-49D3A1/2020\|EPI_ISL_742773\|2020 | ···················· | ···················· | ···················· | ···················· | ···················· | ···················· | |
| Z | 6322 hCoV-19/Wales/PHWC-49D3B0/2020\|EPI_ISL_742774\|2020 | ···················· | ···················· | ···················· | ···················· | ···················· | ···················· | |
| Z | 6323 hCoV-19/Wales/PHWC-49D3CF/2020\|EPI_ISL_742775\|2020 | ···················· | ···················· | ···················· | ···················· | ···················· | ···················· | |
| Z | 6324 hCoV-19/Wales/PHWC-49D3DE/2020\|EPI_ISL_742776\|2020 | ···················· | ···················· | ···················· | ···················· | ···················· | ···················· | |
| Z | 6325 hCoV-19/Wales/PHWC-49D3ED/2020\|EPI_ISL_742777\|2020 | ···················· | ···················· | ···················· | ···················· | ···················· | ···················· | |
| Z | 6326 hCoV-19/Wales/PHWC-49D3FC/2020\|EPI_ISL_742778\|2020 | ···················· | ···················· | ···················· | ···················· | ···················· | ···················· | |
| Z | 6327 hCoV-19/Wales/PHWC-49D408/2020\|EPI_ISL_742779\|2020 | ···················· | ···················· | ···················· | ···················· | ···················· | ···················· | |
| Z | 6328 hCoV-19/Wales/PHWC-49D417/2020\|EPI_ISL_742780\|2020 | ···················· | ···················· | ···················· | ···················· | ···················· | ···················· | |
| Z | 6329 hCoV-19/Wales/PHWC-49D426/2020\|EPI_ISL_742781\|2020 | ···················· | ···················· | ···················· | ···················· | ···················· | ···················· | |
| Z | 6330 hCoV-19/Wales/PHWC-49D435/2020\|EPI_ISL_742782\|2020 | ···················· | ···················· | ···················· | ···················· | ···················· | ···················· | |
| Z | 6331 hCoV-19/Wales/PHWC-49D444/2020\|EPI_ISL_742783\|2020 | ···················· | ···················· | ···················· | ···················· | ···················· | ···················· | |
| Z | 6332 hCoV-19/Wales/PHWC-49D453/2020\|EPI_ISL_742784\|2020 | ···················· | ···················· | ···················· | ···················· | ···················· | ···················· | |
| Z | 6333 hCoV-19/Wales/PHWC-49D462/2020\|EPI_ISL_742785\|2020 | ···················· | ···················· | ···················· | ···················· | ···················· | ···················· | |
| Z | 6334 hCoV-19/Wales/PHWC-49D471/2020\|EPI_ISL_742786\|2020 | ···················· | ···················· | ···················· | ···················· | ···················· | ···················· | |
| Z | 6335 hCoV-19/Wales/PHWC-49D480/2020\|EPI_ISL_742787\|2020 | ········T··········· | ···················· | ···················· | ···················· | ···················· | ···················· | |
| Z | 6336 hCoV-19/Wales/PHWC-49D49F/2020\|EPI_ISL_742788\|2020 | ···················· | ···················· | ···················· | ···················· | NNN················· | ···················· | 1 |
| Z | 6337 hCoV-19/Wales/PHWC-49D4AE/2020\|EPI_ISL_742789\|2020 | ···················· | ···················· | ···················· | ···················· | ···················· | ···················· | |
| Z | 6338 hCoV-19/Wales/PHWC-49D4BD/2020\|EPI_ISL_742790\|2020 | ···················· | ···················· | ···················· | ···················· | ···················· | ···················· | |
| Z | 6339 hCoV-19/Wales/PHWC-49D541/2020\|EPI_ISL_742797\|2020 | ···················· | ···················· | ···················· | ···················· | ···················· | ···················· | |
| Z | 6340 hCoV-19/Wales/PHWC-49D550/2020\|EPI_ISL_742798\|2020 | ···················· | ···················· | ···················· | ···················· | ···················· | ···················· | |
| Z | 6341 hCoV-19/Luxembourg/LNS9890146/2020\|EPI_ISL_745002\| | ···················· | ···················· | ···················· | ···················· | ···················· | ···················· | |
| Z | 6342 hCoV-19/Luxembourg/LNS9897343/2020\|EPI_ISL_745004\| | ···················· | ···················· | ···················· | ···················· | ···················· | ···················· | |
| Z | 6343 hCoV-19/Luxembourg/LNS9902177/2020\|EPI_ISL_745005\| | ···················· | ···················· | ···················· | ···················· | ···················· | ···················· | |
| Z | 6344 hCoV-19/Luxembourg/LNS9903685/2020\|EPI_ISL_745006\| | ···················· | ···················· | ···················· | ···················· | ···················· | ···················· | |
| Z | 6345 hCoV-19/Luxembourg/LNS9905451/2020\|EPI_ISL_745008\| | ···················· | ···················· | ···················· | ···················· | ···················· | ···················· | |
| Z | 6346 hCoV-19/Luxembourg/LNS9905790/2020\|EPI_ISL_745009\| | ···················· | ···················· | ···················· | ···················· | ···················· | ···················· | |
| Z | 6347 hCoV-19/Luxembourg/LNS9916163/2020\|EPI_ISL_745010\| | ···················· | ···················· | ···················· | ···················· | ···················· | ···················· | |

| | | | | | | | |
|---|---|---|---|---|---|---|---|
| Z | 6348 hCoV-19/Luxembourg/LNS9923262/2020\|EPI_ISL_745011\| | ···················· | ···················· | ····················· | ···················· | ···················· | ···················· |
| Z | 6349 hCoV-19/Luxembourg/LNS9926590/2020\|EPI_ISL_745012\| | ···················· | ···················· | ····················· | ···················· | ···················· | ···················· |
| Z | 6350 hCoV-19/Luxembourg/LNS9933159/2020\|EPI_ISL_745013\| | ···················· | ···················· | ····················· | ···················· | ···················· | ···················· |
| Z | 6351 hCoV-19/Luxembourg/LNS9934510/2020\|EPI_ISL_745014\| | ···················· | ···················· | ····················· | ···················· | ···················· | ···················· |
| Z | 6352 hCoV-19/Luxembourg/LNS9940146/2020\|EPI_ISL_745016\| | ···················· | ···················· | ····················· | ···················· | ···················· | ···················· |
| Z | 6353 hCoV-19/Luxembourg/LNS9941347/2020\|EPI_ISL_745017\| | ···················· | ···················· | ····················· | ···················· | ······T············· | ···················· |
| Z | 6354 hCoV-19/Luxembourg/LNS9953002/2020\|EPI_ISL_745018\| | ···················· | ···················· | ····················· | ···················· | ···················· | ···················· |
| Z | 6355 hCoV-19/Luxembourg/LNS9966424/2020\|EPI_ISL_745020\| | ···················· | ···················· | ····················· | ···················· | ···················· | ···················· |
| Z | 6356 hCoV-19/Luxembourg/LNS9967957/2020\|EPI_ISL_745021\| | ···················· | ···················· | ····················· | ···················· | ···················· | ···················· |
| Z | 6357 hCoV-19/Luxembourg/LNS9982064/2020\|EPI_ISL_745022\| | ···················· | ···················· | ····················· | ···················· | ···················· | ···················· |
| Z | 6358 hCoV-19/Luxembourg/LNS9982997/2020\|EPI_ISL_745023\| | ···················· | ···················· | ····················· | ···················· | ···················· | ···················· |
| Z | 6359 hCoV-19/Luxembourg/LNS9989036/2020\|EPI_ISL_745025\| | ···················· | ···················· | ····················· | ···················· | ···················· | ···················· |
| Z | 6360 hCoV-19/Luxembourg/LNS9992190/2020\|EPI_ISL_745026\| | ···················· | ···················· | ····················· | ···················· | ···················· | ···················· |
| Z | 6361 hCoV-19/Luxembourg/LNS9993908/2020\|EPI_ISL_745027\| | ···················· | ···················· | ····················· | ···················· | ···················· | ···················· |
| Z | 6362 hCoV-19/Australia/NSW3725/2020\|EPI_ISL_745029\|2020 | ···················· | ···················· | ····················· | ···················· | ···················· | ···················· |
| Z | 6363 hCoV-19/Australia/NSW3726/2020\|EPI_ISL_745030\|2020 | ···················· | ···················· | ····················· | ···················· | ···················· | ···················· |
| Z | 6364 hCoV-19/Australia/NSW3727/2020\|EPI_ISL_745031\|2020 | ···················· | ···················· | ····················· | ···················· | ···················· | ···················· |
| Z | 6365 hCoV-19/Indonesia/JB-WJHL-ITB-L18952/2020\|EPI_ISL_ | ···················· | ···················· | ····················· | ···················· | ···················· | ···················· |
| Z | 6366 hCoV-19/Israel/CVL-44919/2020\|EPI_ISL_745034\|2020- | ···················· | ···················· | ····················· | ···················· | ···················· | ···················· |
| Z | 6367 hCoV-19/Israel/CVL-45080/2020\|EPI_ISL_745036\|2020- | ···················· | ·A·················· | ····················· | ···················· | ···················· | ···A················ |
| Z | 6369 hCoV-19/Israel/CVL-45176/2020\|EPI_ISL_745038\|2020- | ···················· | ···················· | ····················· | ···················· | ···················· | ···················· |
| Z | 6370 hCoV-19/Israel/CVL-45199/2020\|EPI_ISL_745039\|2020- | ···················· | ···················· | ····················· | ···················· | ···················· | ···················· |
| Z | 6371 hCoV-19/USA/UT-UPHL-2012699605/2020\|EPI_ISL_746300 | ···················· | ·A·················· | ····················· | ···················· | ···················· | ···················· |
| Z | 6372 hCoV-19/USA/UT-UPHL-2012394261/2020\|EPI_ISL_746301 | ···················· | ·A·················· | ····················· | ···················· | ···················· | ···················· |
| Z | 6373 hCoV-19/USA/UT-UPHL-2012662012/2020\|EPI_ISL_746302 | ···················· | ·A·················· | ····················· | ···················· | ···················· | ···················· |
| Z | 6374 hCoV-19/USA/UT-UPHL-2012957226/2020\|EPI_ISL_746303 | ···················· | ···················· | ····················· | ···················· | ···················· | ···················· |
| Z | 6375 hCoV-19/USA/UT-UPHL-2012098996/2020\|EPI_ISL_746304 | ···················· | ·A·················· | ····················· | ···················· | ···················· | ···················· |
| Z | 6376 hCoV-19/USA/UT-UPHL-2012135808/2020\|EPI_ISL_746305 | ···················· | ·A·················· | ····················· | ···················· | ···················· | ···················· |
| Z | 6377 hCoV-19/USA/UT-UPHL-2012974332/2020\|EPI_ISL_746306 | ···················· | ···················· | ····················· | ···················· | ···················· | ···················· |
| Z | 6378 hCoV-19/USA/UT-UPHL-2012563035/2020\|EPI_ISL_746307 | ···················· | ·A·················· | ····················· | ···················· | ···················· | ···················· |
| Z | 6379 hCoV-19/USA/UT-UPHL-2012135058/2020\|EPI_ISL_746308 | ···················· | ···················· | ····················· | ···················· | ···················· | ···················· |
| Z | 6380 hCoV-19/USA/UT-UPHL-2012015753/2020\|EPI_ISL_746309 | ···················· | ···················· | ····················· | ···················· | ···················· | ···················· |
| Z | 6381 hCoV-19/USA/UT-UPHL-2012105029/2020\|EPI_ISL_746310 | ···················· | ···················· | ····················· | ···················· | ···················· | ···················· |
| Z | 6382 hCoV-19/USA/UT-UPHL-2012739013/2020\|EPI_ISL_746311 | ···················· | ·A·················· | ····················· | ···················· | ···················· | ···················· |
| Z | 6383 hCoV-19/USA/UT-UPHL-2012578966/2020\|EPI_ISL_746312 | ···················· | ···················· | ····················· | ···················· | ···················· | ···················· |
| Z | 6384 hCoV-19/USA/UT-UPHL-2012377088/2020\|EPI_ISL_746313 | ···················· | ···················· | ····················· | ···················· | ···················· | ···················· |
| Z | 6385 hCoV-19/USA/UT-UPHL-2012439954/2020\|EPI_ISL_746314 | ···················· | ·A·················· | ····················· | ···················· | ···················· | ···················· |
| Z | 6386 hCoV-19/USA/UT-UPHL-2012882868/2020\|EPI_ISL_746315 | ···················· | ···················· | ····················· | ···················· | ···················· | ···················· |
| Z | 6387 hCoV-19/USA/UT-UPHL-2012740755/2020\|EPI_ISL_746316 | ···················· | ···················· | ····················· | ···················· | ···················· | ···················· |
| Z | 6388 hCoV-19/USA/UT-UPHL-2012154719/2020\|EPI_ISL_746317 | ···················· | ···················· | ····················· | ···················· | ···················· | ···················· |
| Z | 6389 hCoV-19/Bangladesh/JUST-GC56-105/2020\|EPI_ISL_7463 | ···················· | ···················· | ····················· | ···················· | ···················· | ···················· |
| Z | 6390 hCoV-19/USA/UT-UPHL-2012603713/2020\|EPI_ISL_746341 | ···················· | ···················· | ····················· | ···················· | ···················· | ···················· |
| Z | 6391 hCoV-19/USA/UT-UPHL-2012027320/2020\|EPI_ISL_746360 | ···················· | ···················· | ····················· | ···················· | ···················· | ···················· |
| Z | 6392 hCoV-19/USA/UT-UPHL-2012077875/2020\|EPI_ISL_746361 | ···················· | ···················· | ····················· | ···················· | ···················· | ···················· |
| Z | 6393 hCoV-19/USA/UT-UPHL-2012107439/2020\|EPI_ISL_746362 | ···················· | ···················· | ····················· | ···················· | ···················· | ···················· |
| Z | 6394 hCoV-19/USA/UT-UPHL-2012121628/2020\|EPI_ISL_746363 | ···················· | ·A·················· | ····················· | ···················· | ···················· | ···················· |
| Z | 6395 hCoV-19/USA/UT-UPHL-2012191112/2020\|EPI_ISL_746364 | ···················· | ·A·················· | ····················· | ···················· | ···················· | ···················· |
| Z | 6396 hCoV-19/USA/UT-UPHL-2012191595/2020\|EPI_ISL_746365 | ···················· | ···················· | ····················· | ···················· | ···················· | ···················· |
| Z | 6397 hCoV-19/USA/UT-UPHL-2012214535/2020\|EPI_ISL_746366 | ···················· | ···················· | ····················· | ···················· | ···················· | ···················· |
| Z | 6398 hCoV-19/USA/UT-UPHL-2012283416/2020\|EPI_ISL_746367 | ···················· | ···················· | ····················· | ···················· | ···················· | ···················· |
| Z | 6399 hCoV-19/USA/UT-UPHL-2012316867/2020\|EPI_ISL_746368 | ···················· | ···················· | ····················· | ···················· | ···················· | ···················· |
| Z | 6400 hCoV-19/USA/UT-UPHL-2012379693/2020\|EPI_ISL_746369 | ···················· | ·A·················· | ····················· | ···················· | ···················· | ···················· |
| Z | 6401 hCoV-19/England/NORW-222552/2020\|EPI_ISL_741500\|20 | ···················· | ···················· | ····················· | ···················· | ···················· | ···················· |
| Z | 6402 hCoV-19/England/NORW-2225E9/2020\|EPI_ISL_741506\|20 | ···················· | ···················· | ····················· | ···················· | ···················· | ···················· |
| Z | 6403 hCoV-19/England/NORW-222C8A/2020\|EPI_ISL_741510\|20 | ···················· | ···················· | ····················· | ···················· | ···················· | ···················· |
| Z | 6404 hCoV-19/England/NORW-222C99/2020\|EPI_ISL_741511\|20 | ···················· | ·A·················· | ····················· | ···················· | ···················· | ···A················ |
| Z | 6405 hCoV-19/Wales/PHWC-49D57E/2020\|EPI_ISL_742800\|2020 | ···················· | ···················· | ····················· | ···················· | ···················· | ···················· |
| Z | 6406 hCoV-19/Wales/PHWC-49D58D/2020\|EPI_ISL_742801\|2020 | ···················· | ···················· | ····················· | ···················· | ···················· | ···················· |
| Z | 6407 hCoV-19/Wales/PHWC-49D59C/2020\|EPI_ISL_742802\|2020 | ···················· | ···················· | ····················· | ···················· | ···················· | ···················· |
| Z | 6408 hCoV-19/Wales/PHWC-49D5C9/2020\|EPI_ISL_742804\|2020 | ···················· | ···················· | ····················· | ···················· | ···················· | ···················· |
| Z | 6409 hCoV-19/Wales/PHWC-49D5D8/2020\|EPI_ISL_742805\|2020 | ···················· | ···················· | ····················· | ···················· | ···················· | ···················· |
| Z | 6410 hCoV-19/Wales/PHWC-49D5E7/2020\|EPI_ISL_742806\|2020 | ···················· | ···················· | ····················· | ···················· | ···················· | ···················· |
| Z | 6411 hCoV-19/Wales/PHWC-49D5F6/2020\|EPI_ISL_742807\|2020 | ···················· | ···················· | ····················· | ···················· | ···················· | ···················· |
| Z | 6412 hCoV-19/Wales/PHWC-49D602/2020\|EPI_ISL_742808\|2020 | ···················· | ···················· | ····················· | ···················· | ···················· | ···················· |
| Z | 6413 hCoV-19/Wales/PHWC-49D611/2020\|EPI_ISL_742809\|2020 | ···················· | ···················· | ····················· | ···················· | ···················· | ···················· |
| Z | 6414 hCoV-19/Wales/PHWC-49D66C/2020\|EPI_ISL_742814\|2020 | ···················· | ···················· | ····················· | ···················· | ···················· | ···················· |
| Z | 6415 hCoV-19/Wales/PHWC-49D67B/2020\|EPI_ISL_742815\|2020 | ···················· | ···················· | ····················· | ···················· | ···················· | ···················· |
| Z | 6417 hCoV-19/Wales/PHWC-49D6A8/2020\|EPI_ISL_742818\|2020 | ···················· | ···················· | ····················· | ···················· | ···················· | ···················· |
| Z | 6418 hCoV-19/Wales/PHWC-49D6B7/2020\|EPI_ISL_742819\|2020 | ···················· | ···················· | ····················· | ···················· | ···················· | ···················· |
| Z | 6419 hCoV-19/Wales/PHWC-49D6C6/2020\|EPI_ISL_742820\|2020 | ···················· | ···················· | ····················· | ···················· | ···················· | ···················· |
| Z | 6420 hCoV-19/Wales/PHWC-49D6D5/2020\|EPI_ISL_742821\|2020 | ···················· | ···················· | ····················· | ···················· | ···················· | ···················· |
| Z | 6421 hCoV-19/Wales/PHWC-49D71E/2020\|EPI_ISL_742824\|2020 | ···················· | ···················· | ····················· | ···················· | ····T··············· | ···················· |
| Z | 6422 hCoV-19/Wales/PHWC-49D72D/2020\|EPI_ISL_742825\|2020 | ···················· | ···················· | ····················· | ···················· | ····T··············· | ···················· |
| Z | 6423 hCoV-19/Wales/PHWC-49D73C/2020\|EPI_ISL_742826\|2020 | ···················· | ···················· | ····················· | ···················· | ···················· | ···················· |
| Z | 6424 hCoV-19/Wales/PHWC-49D74B/2020\|EPI_ISL_742827\|2020 | ···················· | ···················· | ····················· | ···················· | ···················· | ···················· |
| Z | 6425 hCoV-19/Wales/PHWC-49D769/2020\|EPI_ISL_742829\|2020 | ···················· | ···················· | ····················· | ···················· | ···················· | ···················· |
| Z | 6426 hCoV-19/Wales/PHWC-49D787/2020\|EPI_ISL_742830\|2020 | ···················· | ···················· | ····················· | ···················· | ···················· | ···················· |
| Z | 6427 hCoV-19/Wales/PHWC-49D7A5/2020\|EPI_ISL_742832\|2020 | ···················· | ···················· | ····················· | ···················· | ···················· | ···················· |
| Z | 6428 hCoV-19/Wales/PHWC-49D7B4/2020\|EPI_ISL_742833\|2020 | ···················· | ···················· | ····················· | ···················· | ···················· | ···················· |
| Z | 6431 hCoV-19/Wales/PHWC-49D857/2020\|EPI_ISL_742838\|2020 | ···················· | ···················· | ····················· | ···················· | ···················· | ···················· |

| | | | | | | | |
|---|---|---|---|---|---|---|---|
| Z | 6433 hCoV-19/Wales/PHWC-49D893/2020\|EPI_ISL_742841\|2020 | ···················· | ···················· | ···················· | ···················· | ···················· | ···················· |
| Z | 6434 hCoV-19/Wales/PHWC-49D8A2/2020\|EPI_ISL_742842\|2020 | ···················· | ···················· | ···················· | ···················· | ···················· | ···················· |
| Z | 6435 hCoV-19/Luxembourg/LNS9866409/2020\|EPI_ISL_745000\| | ···················· | ···················· | ···················· | ···················· | ···················· | ···················· |
| Z | 6436 hCoV-19/Luxembourg/LNS9867466/2020\|EPI_ISL_745001\| | ···················· | ···················· | ···················· | ···················· | ···················· | ···················· |
| Z | 6437 hCoV-19/Luxembourg/LNS9905208/2020\|EPI_ISL_745007\| | ···················· | ···················· | ···················· | ···················· | ······T············· | ···················· |
| Z | 6438 hCoV-19/Israel/CVL-45201/2020\|EPI_ISL_745040\|2020- | ···················· | ···················· | ···················· | ···················· | ···················· | ···················· |
| Z | 6439 hCoV-19/Israel/CVL-45243/2020\|EPI_ISL_745041\|2020- | ···················· | ···················· | ···················· | ···················· | ···················· | ···················· |
| Z | 6440 hCoV-19/Israel/CVL-45507/2020\|EPI_ISL_745042\|2020- | ···················· | ···················· | ···················· | ···················· | ···················· | ···················· |
| Z | 6441 hCoV-19/Israel/CVL-45514/2020\|EPI_ISL_745043\|2020- | ···················· | ···················· | ···················· | ···················· | ···················· | ···················· |
| Z | 6442 hCoV-19/Israel/CVL-45517/2020\|EPI_ISL_745044\|2020- | ···················· | ···················· | ···················· | ···················· | ···················· | ···················· |
| Z | 6443 hCoV-19/Israel/CVL-45523/2020\|EPI_ISL_745045\|2020- | ···················· | ···················· | ···················· | ···················· | ···················· | ···················· |
| Z | 6444 hCoV-19/Israel/CVL-45526/2020\|EPI_ISL_745046\|2020- | ···················· | ···················· | ···················· | ···················· | ···················· | ···················· |
| Z | 6445 hCoV-19/Israel/CVL-45583/2020\|EPI_ISL_745048\|2020- | ···················· | ···················· | ···········T········ | ···················· | ···················· | ···················· |
| Z | 6446 hCoV-19/Israel/CVL-45590/2020\|EPI_ISL_745049\|2020- | ···················· | ···················· | ···················· | ···················· | ···················· | ···················· |
| Z | 6447 hCoV-19/Israel/CVL-45646/2020\|EPI_ISL_745051\|2020- | ···················· | ···················· | ···················· | ···················· | ···················· | ···················· |
| Z | 6448 hCoV-19/Israel/CVL-45862/2020\|EPI_ISL_745052\|2020- | ···················· | ···G················ | ···················· | ···················· | ···················· | ···················· |
| Z | 6449 hCoV-19/Israel/CVL-45908/2020\|EPI_ISL_745053\|2020- | ···················· | ···················· | ···················· | ···················· | ···················· | ···················· |
| Z | 6450 hCoV-19/Israel/CVL-45914/2020\|EPI_ISL_745054\|2020- | ···················· | ···················· | ···················· | ···················· | ···················· | ···················· |
| Z | 6451 hCoV-19/Israel/CVL-45925/2020\|EPI_ISL_745056\|2020- | ···················· | ·A·················· | ···················· | ···················· | ···················· | ···················· |
| Z | 6452 hCoV-19/Israel/CVL-45930/2020\|EPI_ISL_745057\|2020- | ···················· | ···················· | ···················· | ···················· | ···················· | ···················· |
| Z | 6453 hCoV-19/Israel/CVL-45936/2020\|EPI_ISL_745058\|2020- | ···················· | ·A·················· | ···················· | ···················· | ···················· | ···················· |
| Z | 6454 hCoV-19/Israel/CVL-46005/2020\|EPI_ISL_745061\|2020- | ···················· | ···················· | ···················· | ···················· | ···················· | ···················· |
| Z | 6455 hCoV-19/Israel/CVL-46019/2020\|EPI_ISL_745062\|2020- | ···················· | ···················· | ···················· | ···················· | ···················· | ···················· |
| Z | 6456 hCoV-19/Israel/CVL-46022/2020\|EPI_ISL_745063\|2020- | ···················· | ···G················ | ···················· | ···················· | ···················· | ···················· |
| Z | 6457 hCoV-19/Israel/CVL-46024/2020\|EPI_ISL_745064\|2020- | ···················· | ···················· | ···················· | ···················· | ···················· | ···················· |
| Z | 6458 hCoV-19/Israel/CVL-46249/2020\|EPI_ISL_745065\|2020- | ···················· | ···················· | ···················· | ···················· | ···················· | ···················· |
| Z | 6459 hCoV-19/Israel/CVL-46255/2020\|EPI_ISL_745066\|2020- | ···················· | ···················· | ···················· | ···················· | ···················· | ···················· |
| Z | 6460 hCoV-19/Israel/CVL-46317/2020\|EPI_ISL_745067\|2020- | ···················· | ···················· | ··C················· | ···················· | ···················· | ···················· |
| Z | 6461 hCoV-19/Israel/CVL-46323/2020\|EPI_ISL_745068\|2020- | ···················· | ···················· | ···················· | ···················· | ···················· | ···················· |
| Z | 6462 hCoV-19/Israel/CVL-46324/2020\|EPI_ISL_745069\|2020- | ···················· | ···················· | ···················· | ···················· | ···················· | ···················· |
| Z | 6463 hCoV-19/Israel/CVL-46381/2020\|EPI_ISL_745070\|2020- | ···················· | ·A·················· | ···················· | ···················· | ···················· | ···················· |
| Z | 6464 hCoV-19/Israel/CVL-46394/2020\|EPI_ISL_745071\|2020- | ···················· | ···G················ | ···················· | ···················· | ···················· | ···················· |
| Z | 6465 hCoV-19/Israel/CVL-46398/2020\|EPI_ISL_745072\|2020- | ···················· | ···G················ | ···················· | ···················· | ···················· | ···················· |
| Z | 6466 hCoV-19/Israel/CVL-46435/2020\|EPI_ISL_745073\|2020- | ···················· | ···················· | ···················· | ···················· | ···················· | ···················· |
| Z | 6467 hCoV-19/Israel/CVL-46466/2020\|EPI_ISL_745075\|2020- | ···················· | ···················· | ···················· | ···················· | ···················· | ···················· |
| Z | 6468 hCoV-19/Israel/CVL-46509/2020\|EPI_ISL_745078\|2020- | ···················· | ···················· | ···················· | ···················· | ···················· | ···················· |
| Z | 6469 hCoV-19/Israel/CVL-46567/2020\|EPI_ISL_745079\|2020- | ···················· | ···················· | ···················· | ···················· | ···················· | ···················· |
| Z | 6470 hCoV-19/Israel/CVL-46584/2020\|EPI_ISL_745080\|2020- | ···················· | ···················· | ···················· | ···················· | ···················· | ···················· |
| Z | 6471 hCoV-19/Israel/CVL-46606/2020\|EPI_ISL_745081\|2020- | ···················· | ···················· | ···················· | ···················· | ···················· | ···················· |
| Z | 6472 hCoV-19/Israel/CVL-46648/2020\|EPI_ISL_745083\|2020- | ···················· | ···················· | ···················· | ···················· | ···················· | ···················· |
| Z | 6473 hCoV-19/Israel/CVL-46672/2020\|EPI_ISL_745084\|2020- | ···················· | ···················· | ···················· | ···················· | ···················· | ···················· |
| Z | 6474 hCoV-19/Israel/CVL-46693/2020\|EPI_ISL_745085\|2020- | ···················· | ···················· | ···················· | ···················· | ···················· | ···················· |
| Z | 6475 hCoV-19/Israel/CVL-46695/2020\|EPI_ISL_745086\|2020- | ···················· | ···················· | ···················· | ···················· | ···················· | ···················· |
| Z | 6476 hCoV-19/Germany/NI-18B6/2020\|EPI_ISL_745090\|2020 | ···················· | ···················· | ···················· | ···················· | ···················· | ···················· |
| Z | 6477 hCoV-19/Australia/NSW1275/2020\|EPI_ISL_745091\|2020 | ···················· | ···················· | ···················· | ···················· | ···················· | ···················· |
| Z | 6478 hCoV-19/Australia/NSW1276/2020\|EPI_ISL_745092\|2020 | ···················· | ···················· | ···················· | ···················· | ···················· | ···················· |
| Z | 6479 hCoV-19/Australia/NSW1278/2020\|EPI_ISL_745093\|2020 | ···················· | ···················· | ···················· | ···················· | ···················· | ···················· |
| Z | 6480 hCoV-19/Australia/NSW1279/2020\|EPI_ISL_745094\|2020 | ···················· | ···················· | ···················· | ···················· | ···················· | ···················· |
| Z | 6481 hCoV-19/Australia/NSW1280/2020\|EPI_ISL_745095\|2020 | ···················· | ···················· | ···················· | ···················· | ···················· | ···················· |
| Z | 6482 hCoV-19/Australia/NSW1281/2020\|EPI_ISL_745096\|2020 | ···················· | ···················· | ···················· | ···················· | ···················· | ···················· |
| Z | 6483 hCoV-19/Australia/NSW1266/2020\|EPI_ISL_745097\|2020 | ···················· | ···················· | ···················· | ···················· | ···················· | ···················· |
| Z | 6484 hCoV-19/Australia/NSW1267/2020\|EPI_ISL_745098\|2020 | ···················· | ···················· | ···················· | ···················· | ···················· | ···················· |
| Z | 6485 hCoV-19/Australia/NSW1268/2020\|EPI_ISL_745099\|2020 | ···················· | ···················· | ···················· | ···················· | ···················· | ···················· |
| Z | 6486 hCoV-19/USA/UT-UPHL-2012393067/2020\|EPI_ISL_746370 | ···················· | ···················· | ···················· | ···················· | ···················· | ···················· |
| Z | 6487 hCoV-19/USA/UT-UPHL-2012465518/2020\|EPI_ISL_746371 | ···················· | ·A·················· | ···················· | ···················· | ···················· | ···················· |
| Z | 6488 hCoV-19/USA/UT-UPHL-2012538966/2020\|EPI_ISL_746372 | ···················· | ···················· | ···················· | ···················· | ···················· | ···················· |
| Z | 6489 hCoV-19/USA/UT-UPHL-2012550080/2020\|EPI_ISL_746373 | ···················· | ···················· | ···················· | ···················· | ···················· | ···················· |
| Z | 6490 hCoV-19/USA/UT-UPHL-2012602043/2020\|EPI_ISL_746374 | ···················· | ···················· | ···················· | ···················· | ···················· | ···················· |
| Z | 6491 hCoV-19/USA/UT-UPHL-2012604380/2020\|EPI_ISL_746375 | ···················· | ···················· | ···················· | ···················· | ···················· | ···················· |
| Z | 6492 hCoV-19/USA/UT-UPHL-2012615779/2020\|EPI_ISL_746376 | ···················· | ···················· | ···················· | ···················· | ···················· | ···················· |
| Z | 6493 hCoV-19/USA/UT-UPHL-2012651600/2020\|EPI_ISL_746377 | ···················· | ···················· | ···················· | ···················· | ···················· | ···················· |
| Z | 6494 hCoV-19/USA/UT-UPHL-2012681522/2020\|EPI_ISL_746378 | ···················· | ···················· | ···················· | ···················· | ···················· | ···················· |
| Z | 6495 hCoV-19/USA/UT-UPHL-2012724265/2020\|EPI_ISL_746379 | ···················· | ·A·················· | ···················· | ···················· | ···················· | ···················· |
| Z | 6496 hCoV-19/USA/UT-UPHL-2012731963/2020\|EPI_ISL_746380 | ···················· | ···················· | ···················· | ···················· | ···················· | ···················· |
| Z | 6497 hCoV-19/USA/UT-UPHL-2012777849/2020\|EPI_ISL_746381 | ···················· | ···················· | ···················· | ···················· | ···················· | ···················· |
| Z | 6498 hCoV-19/USA/UT-UPHL-2012790266/2020\|EPI_ISL_746382 | ···················· | ···················· | ···················· | ···················· | ···················· | ···················· |
| Z | 6499 hCoV-19/USA/UT-UPHL-2012890506/2020\|EPI_ISL_746383 | ···················· | ···················· | ···················· | ···················· | ···················· | ···················· |
| Z | 6500 hCoV-19/USA/UT-UPHL-2012911463/2020\|EPI_ISL_746384 | ···················· | ···················· | ···················· | ···················· | ···················· | ···················· |
| Z | 6501 hCoV-19/USA/UT-UPHL-2012917003/2020\|EPI_ISL_746385 | ···················· | ·A·················· | ···················· | ···················· | ···················· | ···················· |
| Z | 6502 hCoV-19/USA/UT-UPHL-2012945035/2020\|EPI_ISL_746386 | ···················· | ···················· | ···················· | ···················· | ···················· | ···················· |
| Z | 6503 hCoV-19/USA/UT-UPHL-2012984468/2020\|EPI_ISL_746387 | ···················· | ···················· | ···················· | ···················· | ···················· | ···················· |
| Z | 6504 hCoV-19/USA/UT-UPHL-2012997217/2020\|EPI_ISL_746388 | ···················· | ···················· | ···················· | ···················· | ···················· | ···················· |

**Removed from analysis**

| A | B | C | D | E | F | G | H | I | J | K | L |
|---|---|---|---|---|---|---|---|---|---|---|---|
| Database | Database ID | GISAID ID | GCTGCAATCGTGCTACAACT | TGAACTGTTGCGACTACGTG | TTCGGAAGAGACAGGTACGTT | CACACAATCGAAGCGCAGTA | GCTGGTGCTGCAGCTTATTA | AGGGTCAAGTGCACAGTCTA | Incomplete? | Poor quality? | |
| A | 80 | hCoV-19/France/ARA-10968/2020\|EPI_ISL_418431\|2020- | ···················· | ···················· | ···················· | ··········NNNN······ | ···················· | ···················· | | 1 | |
| A | 82 | hCoV-19/Switzerland/BL-UHB-42177236/2020\|EPI_ISL_4 | ···················· | ···················· | ···················· | ···················· | NNNNNNNNNNNNNNNNNNNN | NNNNNNNNNNNNNNNNNNNN | | 1 | |
| A | 136 | hCoV-19/England/20139051704/2020\|EPI_ISL_420702\|20 | ···················· | ···················· | ···················· | ···················· | ·NNN··N········N···· | ···················· | | 1 | |
| A | 139 | hCoV-19/England/20139055204/2020\|EPI_ISL_420714\|20 | ···················· | ···················· | ···················· | ···················· | NNNNNNNNNNNNNNNNNNNN | NNNNNNNNNNNNNNNNNNNN | | 1 | |
| A | 142 | hCoV-19/England/20139057004/2020\|EPI_ISL_420717\|20 | ···················· | ···················· | ···················· | ···················· | ·N···N·N·····N·NN·NN | N·N·N··············· | | 1 | |
| A | 145 | hCoV-19/England/20139057504/2020\|EPI_ISL_420720\|20 | ···················· | ···················· | ···················· | ···················· | NNNNNNNNNNNNNNNNNNNN | ···················· | | 1 | |
| A | 152 | hCoV-19/England/20139059804/2020\|EPI_ISL_420729\|20 | ···················· | ···················· | ···················· | ···················· | NNNNNNNNNNNN···N···· | ···················· | | 1 | |
| A | 156 | hCoV-19/England/20142062904/2020\|EPI_ISL_420775\|20 | ··T················· | ···················· | ···················· | ···················· | NNNNNNNNNNNNNNNNNNNN | ···················· | | 1 | |
| A | 173 | hCoV-19/Australia/NT12/2020\|EPI_ISL_426900\|2020 | ···················· | ···················· | ···················· | ···················· | NNNNNNNNNNNNNNNNNNNN | NNNNNNNNNNNNNNNNNNNN | | 1 | |
| A | 174 | hCoV-19/Australia/NT13/2020\|EPI_ISL_426901\|2020 | ···················· | ···················· | ···················· | ···················· | NNNNNNNNNNNNNNNNNNNN | NNNNNNNNNNNNNNNNNNNN | | 1 | |
| A | 176 | hCoV-19/Australia/VIC473/2020\|EPI_ISL_426906\|2020- | ···················· | ···A················ | ···················· | ···················· | NNNNNNNNNNNNNNNNNNNN | NNNNNNNNNNNNNNNNNNNN | | 1 | |
| A | 177 | hCoV-19/Australia/VIC475/2020\|EPI_ISL_426907\|2020- | ···················· | ···················· | ···················· | ···················· | ·NNNNNN············· | NNNNNNNNNNNNNNN····· | | 1 | |
| A | 178 | hCoV-19/Australia/VIC478/2020\|EPI_ISL_426910\|2020- | ···················· | ···················· | ···················· | ···················· | NNNNNNNNNNNNNNNNNNNN | NNNNNNNNNNNNNNNNNNNN | | 1 | |
| A | 240 | hCoV-19/Thailand/Bangkok-0045/2020\|EPI_ISL_429181\| | ···················· | ···················· | ···················· | ···················· | NNNNNNNNNNNNNNNNNNNN | ···················· | | 1 | |
| A | 316 | hCoV-19/Switzerland/BL-UHB-42177430/2020\|EPI_ISL_4 | ···················· | ···················· | ···················· | ···················· | NNNNNNNNNNNNNNNNNNNN | NNNNNNNNNNNNNNNNNNNN | | 1 | |
| A | 317 | hCoV-19/Switzerland/BL-UHB-42202622/2020\|EPI_ISL_4 | ···················· | ···················· | ···················· | ···················· | NNNNNNNNNNNNNNNNNNNN | NNNNNNNNNNNNNNNNNNNN | | 1 | |
| A | 359 | hCoV-19/England/20139059904/2020\|EPI_ISL_420730\|20 | ···················· | ···················· | ···················· | ···················· | NNNNNNNNNNNNNNNNNNNN | NNNNNNNNNNNNNNNNNNNN | | 1 | |
| A | 366 | hCoV-19/England/20139062504/2020\|EPI_ISL_420741\|20 | ···················· | ···················· | ···················· | ···················· | ··N················· | N·NN·NN············· | | 1 | |
| A | 369 | hCoV-19/England/20139063204/2020\|EPI_ISL_420744\|20 | ···················· | ···················· | ···················· | ···················· | ···················· | NNNNNNNNN··········· | | 1 | |
| A | 386 | hCoV-19/England/20142062204/2020\|EPI_ISL_420773\|20 | ···················· | ···················· | ···················· | ···················· | ·NNNNNNNNNNNNNNNNNNN | NNNNNNNNNNNNNNNNNNNN | | 1 | |
| A | 389 | hCoV-19/USA/WA-UW-1729/2020\|EPI_ISL_424220\|2020-03 | ···················· | ···················· | ···················· | ··········NNNNNNNN·· | ···················· | ···················· | | 1 | |
| A | 483 | hCoV-19/Australia/VIC913/2020\|EPI_ISL_426882\|2020- | ···················· | ···················· | ···················· | ···················· | NNNNNNNNNNNNNNNNNNNN | NNNNNNNNNNNNNNNNNNNN | | 1 | |
| A | 499 | hCoV-19/Australia/NT10/2020\|EPI_ISL_426898\|2020 | ···················· | ···················· | ···················· | ···················· | NNNNNNNNNNNNNNNNNNNN | NNNNNNNNNNNNNNNNNNNN | | 1 | |
| A | 500 | hCoV-19/Australia/NT11/2020\|EPI_ISL_426899\|2020 | ···················· | ···················· | ···················· | ···················· | NNNNNNN············· | ···················· | | 1 | |
| A | 546 | hCoV-19/England/20108006802/2020\|EPI_ISL_417254\|20 | ···················· | ···················· | ···················· | ···················· | NNNNNNNNNNNNNNNNNNNN | NNNNNNNNNNNNNNNNNNNN | | 1 | |
| A | 549 | hCoV-19/England/20108007402/2020\|EPI_ISL_417257\|20 | ···················· | ···················· | ···················· | ···················· | ·NNNNNNNNNNNNNNNNNNN | NNNNNNNNNNNNNNNNNNNN | | 1 | |
| A | 573 | hCoV-19/Germany/BY-V2010486/2020\|EPI_ISL_420898\|20 | ···················· | ···················· | ···················· | ···················· | NNNNNNNNNNNNNNNNNNNN | NNNNNNNNNNNNNNNNNNNN | | 1 | |
| A | 641 | hCoV-19/Australia/VIC551/2020\|EPI_ISL_426813\|2020- | ···················· | ···················· | ···················· | ···················· | NNNNNNN············· | ···················· | | 1 | |
| A | 644 | hCoV-19/Australia/VIC556/2020\|EPI_ISL_426817\|2020- | ···················· | ···················· | ···················· | ···················· | NNNNNNNNNNNNNNNNNNNN | NNNNNNNNNNNNNNNNNNNN | | 1 | |
| A | 648 | hCoV-19/Australia/VIC563/2020\|EPI_ISL_426824\|2020- | ···················· | ···················· | ···················· | ···················· | NNNNNNN············· | ···················· | | 1 | |
| A | 656 | hCoV-19/Australia/VIC574/2020\|EPI_ISL_426833\|2020- | ···················· | ···················· | ···················· | ···················· | NNNNNNNNNNNNNNNNNNNN | NNNNNNNNNNNNNNNNNNNN | | 1 | |
| A | 657 | hCoV-19/Australia/VIC576/2020\|EPI_ISL_426835\|2020- | ···················· | ···················· | ···················· | ···················· | ····N··············· | NNNNNNNNNNNNNNNNNNNN | | 1 | |
| A | 658 | hCoV-19/Australia/VIC578/2020\|EPI_ISL_426836\|2020- | ···················· | ···················· | ···················· | ···················· | NNNNNNNNNNNNNNNNNNNN | NNNNNNNNN··········· | | 1 | |
| A | 659 | hCoV-19/Australia/VIC580/2020\|EPI_ISL_426837\|2020- | ···················· | ···················· | ···················· | ···················· | NNNNNNNNNNNNNNNNNNNN | NNNNNNNNNNNNNNNNNNNN | | 1 | |
| A | 660 | hCoV-19/Australia/VIC581/2020\|EPI_ISL_426838\|2020- | ···················· | ···················· | ···················· | ···················· | NNNNNNNNNNNNNNNNNNNN | NNNNNNNNNNNNNNNNNNNN | | 1 | |
| A | 661 | hCoV-19/Australia/VIC582/2020\|EPI_ISL_426839\|2020- | ···················· | ···················· | ···················· | ···················· | NNNNNNNNNNNNNNNNNNNN | ···················· | | 1 | |
| A | 662 | hCoV-19/Australia/VIC583/2020\|EPI_ISL_426840\|2020- | ···················· | ···················· | ···················· | ···················· | NNNNNNNNNNNNNNNNNNNN | NNNNNNNNNNNNNNNNNNNN | | 1 | |
| A | 664 | hCoV-19/Australia/VIC585/2020\|EPI_ISL_426842\|2020- | ···················· | ···················· | ···················· | ···················· | NNNNNNNNNNNNNNNNNNNN | NNNNNNNNNNNNNNNNNNNN | | 1 | |
| A | 665 | hCoV-19/Australia/VIC587/2020\|EPI_ISL_426844\|2020- | ···················· | ···················· | ···················· | ···················· | NNNNNNNNNNNNNNNNNNNN | NNNNNNNNNNNNNNNNNNNN | | 1 | |
| A | 666 | hCoV-19/Australia/VIC589/2020\|EPI_ISL_426846\|2020- | ···················· | ···················· | ···················· | ···················· | NNNNNNNNNNNNNNNNNNNN | NNNNNNNNNNNNNNNNNNNN | | 1 | |
| A | 667 | hCoV-19/Australia/VIC590/2020\|EPI_ISL_426847\|2020- | ···················· | ···················· | ···················· | ···················· | NNNNNNN············· | ···················· | | 1 | |
| A | 668 | hCoV-19/Australia/VIC591/2020\|EPI_ISL_426848\|2020- | ···················· | ···················· | ···················· | ···················· | NNNNNNNNNNNNNNNNNNNN | ···················· | | 1 | |
| A | 669 | hCoV-19/Australia/VIC593/2020\|EPI_ISL_426850\|2020- | ···················· | ···················· | ···················· | ···················· | ···············NNNNN | ···················· | | 1 | |
| A | 670 | hCoV-19/Australia/VIC594/2020\|EPI_ISL_426851\|2020- | ···················· | ···················· | ···················· | ···················· | NNNNNNNNNNNNNNNNNNNN | NNNNNNNNNNNNNNNNNNNN | | 1 | |
| A | 671 | hCoV-19/Australia/VIC595/2020\|EPI_ISL_426852\|2020- | ···················· | ···················· | ···················· | ···················· | ·NNNNNNNNNNNNNNNNNNN | NNNN················ | | 1 | |
| A | 672 | hCoV-19/Australia/VIC596/2020\|EPI_ISL_426853\|2020- | ···················· | ···················· | ···················· | ···················· | NNNNNNNNNNNNNNNNNNNN | NNNNNNNNNNNNNNNNNNNN | | 1 | |
| A | 679 | hCoV-19/Australia/VIC607/2020\|EPI_ISL_426862\|2020- | ···················· | ···················· | ···················· | ···················· | NNNNNNNNNNNNNNNNNNNN | NNNNNNNNNNNNNNNNNNNN | | 1 | |
| A | 680 | hCoV-19/Australia/VIC610/2020\|EPI_ISL_426864\|2020- | ···················· | ···················· | ···················· | ···················· | NNNNNNNNNNNN········ | NNNNNNNNNNNNNNNN···· | | 1 | |
| A | 681 | hCoV-19/Australia/VIC611/2020\|EPI_ISL_426865\|2020- | ···················· | ···················· | ···················· | ···················· | NNNNNNN············· | ···················· | | 1 | |
| A | 683 | hCoV-19/Australia/VIC615/2020\|EPI_ISL_426870\|2020- | ···················· | ···················· | ···················· | ···················· | NNNNNNNNNNNNNNNNNNNN | ···················· | | 1 | |
| A | 685 | hCoV-19/Australia/VIC629/2020\|EPI_ISL_426873\|2020- | ···················· | ···················· | ···················· | ···················· | ···················· | NNNNNNNNNNNNNNNNNNNN | | 1 | |
| A | 687 | hCoV-19/Australia/VIC633/2020\|EPI_ISL_426875\|2020- | ···················· | ···················· | ···················· | ···················· | ···················· | NNNNNNNNNNNNNNNNNNNN | | 1 | |
| A | 688 | hCoV-19/Australia/VIC635/2020\|EPI_ISL_426876\|2020- | ···················· | ···················· | ···················· | ···················· | NNNNNNNNNNNNNNNNNNNN | ···················· | | 1 | |
| A | 690 | hCoV-19/Australia/VIC701/2020\|EPI_ISL_426878\|2020- | ···················· | ···················· | ···················· | ···················· | NNNNNNNNNNNNNNNNNNNN | NNNNNNNNNNNNNNNNNNNN | | 1 | |
| A | 716 | hCoV-19/DRC/998/2020\|EPI_ISL_429254\|2020-04-03 | ···················· | ···················· | ···················· | ···················· | NNNNNNNNNNNNNNNNNNNN | NNNNNNNNNNNNNNN····· | | 1 | |
| A | 719 | hCoV-19/DRC/1234/2020\|EPI_ISL_429258\|2020-04-06 | ···················· | ···················· | ···················· | ···················· | NNNNNNNNNNNNNNNNNNNN | N··················· | | 1 | |
| A | 731 | hCoV-19/USA/UT-0514/2020\|EPI_ISL_430060\|2020-04-03 | ···················· | ···················· | ···················· | ···················· | ···················· | NNNNNNNNNNNNNNN····· | | 1 | |
| A | 791 | hCoV-19/England/20102087902/2020\|EPI_ISL_417215\|20 | ···················· | ···················· | ···················· | ···················· | ···················· | ·NN·····N·N····N···· | | 1 | |
| A | 792 | hCoV-19/England/20102098802/2020\|EPI_ISL_417217\|20 | ···················· | ···················· | ···················· | ···················· | ···················· | NNNNNNNNNNNNNNNNNNNN | | 1 | |
| A | 953 | hCoV-19/England/20134017404/2020\|EPI_ISL_423094\|20 | ···················· | ···················· | ···················· | ···················· | NNNNNNNNNNNNNNNNNNNN | NNNNNNNNNNNNNNNNNNNN | | 1 | |
| A | 957 | hCoV-19/England/20134018402/2020\|EPI_ISL_423098\|20 | ···················· | ···················· | ···················· | ···················· | NNNNNNNNNNNNNNNNNNNN | NNNNNNNNNNNNNNN····· | | 1 | |
| A | 1109 | hCoV-19/Thailand/Bangkok-0017/2020\|EPI_ISL_423039\| | ···················· | ···················· | ···················· | ···················· | NNNNNNNNNNNNNNNNNNNN | NNNNNNNNNNNNNNNNNNNN | | 1 | |
| A | 1110 | hCoV-19/Thailand/Bangkok-0019/2020\|EPI_ISL_423040\| | ···················· | ···················· | ···················· | ···················· | NNNNNNNNNNNNNNNNNNNN | ···················· | | 1 | |
| A | 1119 | hCoV-19/England/20134003004/2020\|EPI_ISL_423055\|20 | ···················· | ···················· | ···················· | ···················· | NNNNNNNNNNNNNNNNNNNN | NNNNNNNNNNNNNNNNNNNN | | 1 | |
| A | 1120 | hCoV-19/England/20134003104/2020\|EPI_ISL_423056\|20 | ···················· | ···················· | ···················· | ···················· | ·NNNNNNNNNNNNNNNNNNN | ···················· | | 1 | |
| A | 1123 | hCoV-19/England/20134003404/2020\|EPI_ISL_423059\|20 | ···················· | ···················· | ···················· | ···················· | NNNNNNNNNN·NNNN····N | ··N················· | | 1 | |
| A | 1129 | hCoV-19/England/20134006404/2020\|EPI_ISL_423069\|20 | ···················· | ···················· | ···················· | ···················· | ·NNNNNNNNNNNNNNNNNNN | NNNNN·NNNNNNNNNNNNN· | | 1 | |
| A | 1143 | hCoV-19/England/20134015902/2020\|EPI_ISL_423087\|20 | ···················· | ···················· | ···················· | ···················· | ·NNNNNNNNNNNNNNNNNNN | NNN·N·············N· | | 1 | |
| A | 1242 | hCoV-19/Australia/VIC483/2020\|EPI_ISL_426914\|2020- | ···················· | ···················· | ···················· | ···················· | NNNNNNNNNNNNNNNNNNNN | NNNNNNNNNNNNNNNNNNNN | | 1 | |

| | | | | | | | | |
|---|---|---|---|---|---|---|---|---|
| A | 1243 hCoV-19/Australia/VIC486/2020\|EPI_ISL_426917\|2020- | ···················· | ···················· | ···················· | ···················· | NNNNNNNNNNNNNNNNNNNN | NNNNNNNNNNNNNNNNNNNN | 1 |
| A | 1244 hCoV-19/Australia/VIC487/2020\|EPI_ISL_426918\|2020- | ···················· | ···················· | ···················· | ···················· | NNNNNNNNNNNNNNNNNNNN | NNNNNNNNNNNNNNNNNNNN | 1 |
| A | 1245 hCoV-19/Australia/VIC488/2020\|EPI_ISL_426919\|2020- | ···················· | ···················· | ···················· | ···················· | NNNNNNNNNNNNNNNNNNNN | NNNNNNNNNNNNNNNNNNNN | 1 |
| A | 1246 hCoV-19/Australia/VIC489/2020\|EPI_ISL_426920\|2020- | ···················· | ···················· | ···················· | ···················· | ···················· | NNNNNNNNNNNNNNNNNNNN | 1 |
| A | 1247 hCoV-19/Australia/VIC490/2020\|EPI_ISL_426921\|2020- | ···················· | ···················· | ···················· | ···················· | NNNNNNNNNNNNNNNNNNNN | NNNNNNNNNNNNNNNNNNNN | 1 |
| A | 1248 hCoV-19/Australia/VIC491/2020\|EPI_ISL_426922\|2020- | ···················· | ···················· | ···················· | ···················· | NNNNNNNNNNNNNNNNNNNN | ···················· | 1 |
| A | 1249 hCoV-19/Australia/VIC619/2020\|EPI_ISL_426923\|2020- | ···················· | ···················· | ···················· | ···················· | NNNNNNNNNNNNNNNNNNNN | NNNNNNNNNNNNNNNNNNNN | 1 |
| A | 1250 hCoV-19/Australia/VIC620/2020\|EPI_ISL_426924\|2020- | ···················· | ···················· | ···················· | ···················· | NNNNNNNNNNNNNNNNNNNN | NNNNNNNNNNNNNNNNNNNN | 1 |
| A | 1251 hCoV-19/Australia/VIC622/2020\|EPI_ISL_426926\|2020- | ···················· | ···················· | ···················· | ···················· | NNNNNNNNNNNNNNNNNNNN | NNNNNNNNNNNNNNNNNNNN | 1 |
| A | 1253 hCoV-19/Australia/VIC625/2020\|EPI_ISL_426929\|2020- | ···················· | ···················· | ···················· | ···················· | NNNNNNNNNNNNNNNNNNNN | ···················· | 1 |
| A | 1254 hCoV-19/Australia/VIC626/2020\|EPI_ISL_426930\|2020- | ···················· | ···················· | ···················· | ···················· | ···················· | NNNNNNNNNNNNNNN····· | 1 |
| A | 1255 hCoV-19/Australia/VIC627/2020\|EPI_ISL_426931\|2020- | ···················· | ···················· | ···················· | ···················· | NNNNNNN············· | ···················· | 1 |
| A | 1257 hCoV-19/Australia/VIC637/2020\|EPI_ISL_426933\|2020- | ···················· | ···················· | ···················· | ···················· | NNNNNNNNNNNNNNNNNNNN | NNNNNNNNNNNNNNNNNNNN | 1 |
| A | 1264 hCoV-19/Australia/VIC651/2020\|EPI_ISL_426947\|2020- | ···················· | ···················· | ···················· | ···················· | ···················· | NNNN················ | 1 |
| A | 1266 hCoV-19/Australia/VIC655/2020\|EPI_ISL_426951\|2020- | ···················· | ···················· | ···················· | ···················· | NNNNNNNNNNNN········ | ···················· | 1 |
| A | 1268 hCoV-19/Australia/VIC657/2020\|EPI_ISL_426953\|2020- | ···················· | ···················· | ···················· | ···················· | NNNNNNNNNNNNNNNNNNNN | ···················· | 1 |
| A | 1269 hCoV-19/Australia/VIC658/2020\|EPI_ISL_426954\|2020- | ···················· | ···················· | ···················· | ···················· | NNNNNNNNNNNNNNNNNNNN | ···················· | 1 |
| A | 1270 hCoV-19/Australia/VIC659/2020\|EPI_ISL_426955\|2020- | ···················· | ···················· | ···················· | ···················· | ·····NNNNNNNNNNNNNNN | ·················NNN | 1 |
| A | 1271 hCoV-19/Australia/VIC683/2020\|EPI_ISL_426978\|2020- | ···················· | ···················· | ···················· | ···················· | NNNNNNNNNNNNNNNNNNNN | NNNNNNNNNNNNNNNNNNNN | 1 |
| A | 1273 hCoV-19/Australia/VIC687/2020\|EPI_ISL_426981\|2020- | ···················· | ···················· | ···················· | ···················· | NNNNNNNNNNNNNNNNNNNN | NNNNNNNNNNNNNNNNNNNN | 1 |
| A | 1274 hCoV-19/Australia/VIC693/2020\|EPI_ISL_426985\|2020- | ···················· | ···················· | ···················· | ···················· | NNNNNNNNNNNNNNNNNNNN | NNNNNNNNN··········· | 1 |
| A | 1326 hCoV-19/Italy/ABR-IZSGC-TE4959/2020\|EPI_ISL_418259 | ···················· | ···················· | NNNNNNNNNNNNNNNNNNNN | ···················· | ···················· | ···················· | 1 |
| A | 1335 hCoV-19/Switzerland/BS-UHB-42169171/2020\|EPI_ISL_4 | ···················· | ···················· | ···················· | ···················· | NNNNNNNNNNNNNNNNNNNN | NNNNNNNNNNNNNNNNNNNN | 1 |
| A | 1336 hCoV-19/Switzerland/BL-UHB-42174724/2020\|EPI_ISL_4 | ···················· | ···················· | ···················· | ···················· | NNNNNNNNNNNNNNNNNNNN | NNNNNNNNNNNNNNNNNNNN | 1 |
| A | 1337 hCoV-19/Switzerland/BL-UHB-42175213/2020\|EPI_ISL_4 | ···················· | ···················· | ···················· | ···················· | NNNNNNNNNNNNNNNNNNNN | NNNNNNNNNNNNNNNNNNNN | 1 |
| A | 1443 hCoV-19/England/20139009604/2020\|EPI_ISL_421869\|20 | ···················· | ···················· | ···················· | ···················· | ·NNNNNNNNNNNNNNNNNNN | NNNNNNNNNNNNNNNNNNNN | 1 |
| A | 1444 hCoV-19/England/20139009704/2020\|EPI_ISL_421870\|20 | ···················· | ···················· | ···················· | ···················· | ·NNNNNNNNNNNNNNNNNNN | NNNNNNNNNNNNNNNNNNNN | 1 |
| A | 1446 hCoV-19/England/20139011304/2020\|EPI_ISL_421875\|20 | ···················· | ···················· | ···················· | ···················· | NNNNN··N··N·N······· | ···················· | 1 |
| A | 1447 hCoV-19/England/20139014404/2020\|EPI_ISL_421877\|20 | ···················· | ···················· | ···················· | ···················· | ·NNNNNNNNNNNNNNNNNNN | ·········N·········· | 1 |
| A | 1456 hCoV-19/England/20139023604/2020\|EPI_ISL_421888\|20 | ···················· | ···················· | ···················· | ···················· | ·NNNNNNNNNNNNNNNNNNN | ···················· | 1 |
| A | 1459 hCoV-19/England/20139024204/2020\|EPI_ISL_421891\|20 | ···················· | ···················· | ···················· | ···················· | NNNNNNNNNNNNNNNNNNNN | NNNNNNNNNNNNNNNNNNNN | 1 |
| A | 1460 hCoV-19/England/20139024304/2020\|EPI_ISL_421892\|20 | ···················· | ···················· | ···················· | ···················· | ··NNNNNNNNNNNNNNNNNN | NNNNNNNNNNNNNNNNNNNN | 1 |
| A | 1461 hCoV-19/England/20139024604/2020\|EPI_ISL_421893\|20 | ···················· | ···················· | ···················· | ···················· | ·NNNNNNNNNNNNNNNNNNN | NNNNNNNNNNNNNNNNNNNN | 1 |
| A | 1463 hCoV-19/England/20139024904/2020\|EPI_ISL_421895\|20 | ···················· | ···················· | ···················· | ···················· | ··NN···NN·NN········ | ····N··············· | 1 |
| A | 1513 hCoV-19/Australia/VIC660/2020\|EPI_ISL_426956\|2020- | ···················· | ···················· | ···················· | ···················· | NNNNNNN············· | ···················· | 1 |
| A | 1516 hCoV-19/Australia/VIC663/2020\|EPI_ISL_426959\|2020- | ···················· | ···················· | ···················· | ···················· | NNNNNNNNNNNNNNNNNNNN | ····W··············· | 1 |
| A | 1519 hCoV-19/Australia/VIC666/2020\|EPI_ISL_426962\|2020- | ···················· | ·A·················· | ···················· | ···················· | NNNNNNN············· | ···················· | 1 |
| A | 1524 hCoV-19/Australia/VIC674/2020\|EPI_ISL_426970\|2020- | ···················· | ···················· | ···················· | ···················· | ···················· | NNNNNNNNNNNNNNNNNNNN | 1 |
| A | 1527 hCoV-19/Australia/VIC678/2020\|EPI_ISL_426973\|2020- | ···················· | ···················· | ···················· | ···················· | NNNNNNNNNNNN········ | ···················· | 1 |
| A | 1532 hCoV-19/Australia/VIC706/2020\|EPI_ISL_426993\|2020- | ···················· | ···················· | ···················· | ···················· | ···················· | NNNNNNNNNNNNNNNNNNNN | 1 |
| A | 1533 hCoV-19/Australia/VIC707/2020\|EPI_ISL_426994\|2020- | ···················· | ···················· | ···················· | ···················· | NNNNNNNNNNNNNNNNNNNN | NNNNNNNNNNNNNNNNNNNN | 1 |
| A | 1534 hCoV-19/Australia/VIC708/2020\|EPI_ISL_426995\|2020- | ···················· | ···················· | ···················· | ···················· | NNNNNNNNNNNNNNNNNNNN | ···················· | 1 |
| A | 1583 hCoV-19/Switzerland/BL-UHB-42175220/2020\|EPI_ISL_4 | ···················· | ···················· | ···················· | ···················· | NNNNNNNNNNNNNNNNNNNN | NNNNNNNNNNNNNNNNNNNN | 1 |
| A | 1584 hCoV-19/Switzerland/BL-UHB-42176229/2020\|EPI_ISL_4 | ···················· | ···················· | ···················· | ···················· | NNNNNNNNNNNNNNNNNNNN | NNNNNNNNNNNNNNNNNNNN | 1 |
| A | 1629 hCoV-19/England/201380069/2020\|EPI_ISL_421817\|2020 | ···················· | ···················· | ···················· | ···················· | ···················· | ···N·····N···NNNNNNN | 1 |
| A | 1636 hCoV-19/England/201380277/2020\|EPI_ISL_421827\|2020 | ···················· | ···················· | ···················· | ···················· | NNNNNNNNNNNNNNNNNNNN | NNNNNNNNNNNNNNNNNNNN | 1 |
| A | 1638 hCoV-19/England/201380281/2020\|EPI_ISL_421829\|2020 | ···················· | ···················· | ···················· | ···················· | ···················· | NNNNNNNNNNNNNNN·N··N | 1 |
| A | 1639 hCoV-19/England/201380282/2020\|EPI_ISL_421830\|2020 | ···················· | ···················· | ···················· | ···················· | ·NNNNNNNNNNNNNNNNNNN | NNNNNNNNNNNNNNNNNNNN | 1 |
| A | 1640 hCoV-19/England/201380283/2020\|EPI_ISL_421831\|2020 | ···················· | ···················· | ···················· | ···················· | ··NNNNNNNNNNNNNNNNNN | NNNNNNNNNNNNNNN···N· | 1 |
| A | 1645 hCoV-19/England/20138086604/2020\|EPI_ISL_421841\|20 | ···················· | ···················· | ···················· | ···················· | ···················· | NNNNN··············· | 1 |
| A | 1654 hCoV-19/England/20129016104/2020\|EPI_ISL_424001\|20 | ···················· | ···················· | ···················· | ···················· | ··N···N···········NN | ···················· | 1 |
| A | 1656 hCoV-19/England/20129018604/2020\|EPI_ISL_424006\|20 | ···················· | ···················· | ···················· | ···················· | ··N··NN···NNNNNNN·NN | NNNNNNNNNNNNNNNNNNNN | 1 |
| A | 1661 hCoV-19/England/20129023104/2020\|EPI_ISL_424015\|20 | ···················· | ···················· | ···················· | ···················· | ···················· | NNNNNNN············· | 1 |
| A | 1663 hCoV-19/England/20129031804/2020\|EPI_ISL_424019\|20 | ···················· | ···················· | ···················· | ···················· | ··NNNNNNNNNNNNNNNNNN | NNNNNNNNNNNNNNNNNNNN | 1 |
| A | 1665 hCoV-19/England/20129032404/2020\|EPI_ISL_424021\|20 | ···················· | ···················· | ···················· | ···················· | ····NNNNNNNNNNNNNNNN | NNNNNNNNNNNNNNN····· | 1 |
| A | 1676 hCoV-19/England/20129056804/2020\|EPI_ISL_424052\|20 | ···················· | ···················· | ···················· | ···················· | ···················· | NNNNNNNNNNNNNNN····· | 1 |
| A | 1678 hCoV-19/England/20129063804/2020\|EPI_ISL_424062\|20 | ···················· | ···················· | ···················· | ···················· | NNNNNNNNNNNNNNNNNNNN | NNNNNNNNNNNNNNNNNNNN | 1 |
| A | 1683 hCoV-19/England/CAMB-74562/2020\|EPI_ISL_425306\|202 | ···················· | ···················· | ···················· | ···················· | NNNNNNNNNNNNNNNNNNNN | NNNNNNNNNNNNNNNNNNNN | 1 |
| A | 1697 hCoV-19/England/CAMB-7468D/2020\|EPI_ISL_425321\|202 | ···················· | ···················· | ···················· | ···················· | NNNNNNNNNNNNNNNNNNNN | NNNNNNNNNNNNNNNNNNNN | 1 |
| A | 1698 hCoV-19/England/CAMB-7469C/2020\|EPI_ISL_425322\|202 | ···················· | ···················· | ···················· | ···················· | NNNNNNNNNNNNNNNNNNNN | NNNNNNNNNNNNNNNNNNNN | 1 |
| A | 1699 hCoV-19/England/CAMB-746AB/2020\|EPI_ISL_425323\|202 | ···················· | ···················· | ···················· | ···················· | ··············N····· | NN····NN············ | 1 |
| A | 1710 hCoV-19/England/CAMB-74A09/2020\|EPI_ISL_425342\|202 | ···················· | ···················· | ···················· | ···················· | NNNNNNNNNNNNNNNNNNNN | NNNNNNNNNNNNNNNNNNNN | 1 |
| A | 1712 hCoV-19/England/CAMB-74B9D/2020\|EPI_ISL_425351\|202 | ···················· | ···················· | ···················· | ···················· | NNNNNNNNNNNNNNNNNNNN | NNNNNNNNNNNNNNNNNNNN | 1 |
| A | 1713 hCoV-19/England/CAMB-74BAC/2020\|EPI_ISL_425352\|202 | ···················· | ···················· | ···················· | ···················· | NNNNNNNNNNNNNNNNNNNN | NNNNNNNNNNNNNNNNNNNN | 1 |
| A | 1714 hCoV-19/England/CAMB-74C21/2020\|EPI_ISL_425358\|202 | ···················· | ···················· | ···················· | ···················· | NNNNNNNNNNNNNNNNNNNN | NNNNNNNNNNNNNNNNNNNN | 1 |
| A | 1723 hCoV-19/England/CAMB-75464/2020\|EPI_ISL_425390\|202 | ···················· | ···················· | ···················· | ···················· | NNNNNNNNNNNNNNNNNNNN | NNNNNNNNNNNNNNNNNNNN | 1 |
| A | 1724 hCoV-19/England/CAMB-75482/2020\|EPI_ISL_425392\|202 | ···················· | ···················· | ···················· | ···················· | NNNNNNNNNNNNNNNNNNNN | NNNNNNNNNNNNNNNNNNNN | 1 |
| A | 1725 hCoV-19/England/CAMB-75491/2020\|EPI_ISL_425393\|202 | ···················· | ···················· | ···················· | ···················· | NNNNNNNNNNNNNNNNNNNN | NNNNNNNNNNNNNNNNNNNN | 1 |
| A | 1726 hCoV-19/England/CAMB-754B7/2020\|EPI_ISL_425395\|202 | ···················· | ···················· | ···················· | ···················· | NNNNNNNNNNNNNNNNNNNN | NNNNNNNNNNNNNNNNNNNN | 1 |
| A | 1727 hCoV-19/England/CAMB-754EC/2020\|EPI_ISL_425398\|202 | ···················· | ···················· | ···················· | ···················· | NNNNNNNNNNNNNNNNNNNN | NNNNNNNNNNNNNNNNNNNN | 1 |
| A | 1828 hCoV-19/Belgium/ULG-3843/2020\|EPI_ISL_417012\|2020- | ···················· | ···················· | ···················· | ···················· | NNNNNNNNNNNNNNNNNNNN | NNNNNNNNNNNNNNNNNNNN | 1 |
| A | 1829 hCoV-19/Belgium/ULG-4163/2020\|EPI_ISL_417013\|2020- | ···················· | ···················· | ···················· | ···················· | NNNNNNNNNNNNNNNNNNNN | NNNNNNNNNNNNNNNNNNNN | 1 |
| A | 1943 hCoV-19/England/20139044504/2020\|EPI_ISL_420683\|20 | ···················· | ···················· | ···················· | ···················· | NNNNNNNNNNNNNNNNNNNN | ···················· | 1 |
| A | 1944 hCoV-19/England/20139045804/2020\|EPI_ISL_420686\|20 | ···················· | ···················· | ···················· | ···················· | NNNNNNNNNNNNNNNNNNNN | NNNNNNNNNNNNNNNNNNNN | 1 |
| A | 1946 hCoV-19/England/20139046504/2020\|EPI_ISL_420688\|20 | ···················· | ···················· | ···················· | ···················· | NNNNNNNNNNNNN······N | ···················· | 1 |
| A | 1947 hCoV-19/England/20139046604/2020\|EPI_ISL_420689\|20 | ···················· | ···················· | ···················· | ···················· | NNNNNN·NNNN········· | ···················· | 1 |
| A | 1948 hCoV-19/England/20139046704/2020\|EPI_ISL_420690\|20 | ···················· | ···················· | ···················· | ···················· | NNNN················ | ···················· | 1 |
| A | 1957 hCoV-19/England/CAMB-74517/2020\|EPI_ISL_425301\|202 | ···················· | ···················· | ···················· | ···················· | NNNNNNNNNNNNNNNNNNNN | NNNNNNNNNNNNNNNNNNNN | 1 |
| A | 1958 hCoV-19/England/CAMB-74535/2020\|EPI_ISL_425303\|202 | ···················· | ···················· | ···················· | ···················· | NNNNNNNNNNNNNNNNNNNN | NNNNNNNNNNNNNNNNNNNN | 1 |
| A | 1976 hCoV-19/Australia/NT08/2020\|EPI_ISL_426634\|2020 | ···················· | ···················· | ···················· | ···················· | NNNNNNNNNNNNNNNNNNNN | NNNNNNNNNNNNNNNNNNNN | 1 |

| | | | | | | | | |
|---|---|---|---|---|---|---|---|---|
| A | 1977 hCoV-19/Australia/NT18/2020\|EPI_ISL_426635\|2020 | ···················· | ···················· | ···················· | ···················· | ···················· | NNNNNNNNN··········· | 1 |
| A | 2047 hCoV-19/USA/UT-0508/2020\|EPI_ISL_430056\|2020-04-02 | ···················· | ···················· | ···················· | ···················· | NNNNNNNNNNNNNNNNNNNN | NNNNNNNNNNNNNNNNNNNN | 1 |
| A | 2056 hCoV-19/Belgium/ULG-3163/2020\|EPI_ISL_417006\|2020- | ···················· | ···················· | ···················· | ···················· | ··············N····· | NN·N···NN··········· | 1 |
| A | 2152 hCoV-19/France/ARA-13074/2020\|EPI_ISL_420622\|2020- | ···················· | ···················· | ···················· | NNNNNNNNNNNNNNNNNNNN | ···················· | ···················· | 1 |
| A | 2168 hCoV-19/England/20139027304/2020\|EPI_ISL_421903\|20 | ···················· | ···················· | ···················· | ···················· | ·NNNNNNNNNNNNNNNNNNN | NNNNNNNNNNNNNNNNNNNN | 1 |
| A | 2183 hCoV-19/England/20139033304/2020\|EPI_ISL_421920\|20 | ···················· | ···················· | ···················· | ···················· | ·NNNNNNNNNNNNNNNNNNN | NNNNNNNNNNNNNNNNNNNN | 1 |
| A | 2186 hCoV-19/England/20140007202/2020\|EPI_ISL_421924\|20 | ···················· | ···················· | ···················· | ···················· | ··N················· | ··NN····NNNNNN······ | 1 |
| A | 2187 hCoV-19/England/20140008402/2020\|EPI_ISL_421929\|20 | ···················· | ·········A·········· | ···················· | ···················· | ····NNNNNNNNNNNNNNNN | NNNNNNNNNNNNNNNNNNNN | 1 |
| A | 2192 hCoV-19/England/20144014704/2020\|EPI_ISL_421936\|20 | ···················· | ···················· | ···················· | ···················· | ···················· | NNNNNNNNNNNNNNNNNNNN | 1 |
| A | 2198 hCoV-19/England/20144035904/2020\|EPI_ISL_421945\|20 | ···················· | ···················· | ···················· | ···················· | ·NNNNNNNNNNNNNNNNNNN | NNNNNNNNNNNNNNNNNNNN | 1 |
| A | 2200 hCoV-19/England/20144038204/2020\|EPI_ISL_421947\|20 | ···················· | ···················· | ···················· | ···················· | ···················· | N····NNNNN···NN····· | 1 |
| A | 2204 hCoV-19/England/20144065604/2020\|EPI_ISL_421956\|20 | ···················· | ···················· | ···················· | ···················· | ·NNNNNNNNNNNNNNNNNNN | NNNNNNNNNNNNNNNNNNNN | 1 |
| A | 2207 hCoV-19/England/20144072804/2020\|EPI_ISL_421962\|20 | ···················· | ···················· | ···················· | ···················· | ···················· | NNNNNNN············· | 1 |
| A | 2208 hCoV-19/England/20144074804/2020\|EPI_ISL_421963\|20 | ···················· | ···················· | ···················· | ···················· | ·NNNNNNNNNNNNNNNNNNN | NNNNNNNNNNNNNNNNNNNN | 1 |
| A | 2211 hCoV-19/England/20146009204/2020\|EPI_ISL_421976\|20 | ···················· | ···················· | ···················· | ···················· | ···················· | NNNNNNNNN·····N····· | 1 |
| A | 2216 hCoV-19/England/20129147504/2020\|EPI_ISL_424115\|20 | ···················· | ···················· | ···················· | ···················· | NNNNNNNNNNNNNNNNNNNN | NNNNNNNNNNNNNNNNNNNN | 1 |
| A | 2240 hCoV-19/England/CAMB-76E38/2020\|EPI_ISL_425432\|202 | ···················· | ···················· | ···················· | ···················· | NNNNNNNNNNNNNNNNNNNN | NNNNNNNNNNNNNNNNNNNN | 1 |
| A | 2252 hCoV-19/Australia/VIC511/2020\|EPI_ISL_426781\|2020- | ···················· | ···················· | ···················· | ···················· | NNNNNNN············· | ··R················· | 1 |
| A | 2257 hCoV-19/Australia/VIC518/2020\|EPI_ISL_426788\|2020- | ···················· | ···················· | ···················· | ···················· | NNNNNNN············· | ···················· | 1 |
| A | 2260 hCoV-19/Australia/VIC525/2020\|EPI_ISL_426792\|2020- | ···················· | ···················· | ···················· | ···················· | NN····N············· | NNNN················ | 1 |
| A | 2265 hCoV-19/Australia/VIC533/2020\|EPI_ISL_426799\|2020- | ···················· | ···················· | ···················· | ···················· | ·············NNNNNNN | ·················M·· | 1 |
| A | 2299 hCoV-19/USA/UT-085/2020\|EPI_ISL_430034\|2020-03-17 | ···················· | ···················· | ···················· | ···················· | NNNNNNN············· | ···················· | 1 |
| A | 2369 hCoV-19/England/20129158804/2020\|EPI_ISL_424119\|20 | ···················· | ···················· | ···················· | ···················· | NNNNNNNNNNNNNNNNNNNN | NNNNNNNNNNNNNNNNNNNN | 1 |
| A | 2388 hCoV-19/England/CAMB-75516/2020\|EPI_ISL_425401\|202 | ···················· | ···················· | ···················· | ···················· | NNNNNNNNNNNNNNNNNNNN | NNNNNNNNNNNNNNNNNNNN | 1 |
| A | 2389 hCoV-19/England/CAMB-75525/2020\|EPI_ISL_425402\|202 | ···················· | ···················· | ···················· | ···················· | NNNNNNNNNNNNNNNNNNNN | NNNNNNNNNNNNNNNNNNNN | 1 |
| A | 2390 hCoV-19/England/CAMB-75534/2020\|EPI_ISL_425403\|202 | ···················· | ···················· | ···················· | ···················· | NNNNNNNNNNNNNNNNNNNN | NNNNNNNNNNNNNNNNNNNN | 1 |
| A | 2391 hCoV-19/England/CAMB-75543/2020\|EPI_ISL_425404\|202 | ···················· | ···················· | ···················· | ···················· | NNNNNNNNNNNNNNNNNNNN | NNNNNNNNNNNNNNNNNNNN | 1 |
| A | 2392 hCoV-19/England/CAMB-75552/2020\|EPI_ISL_425405\|202 | ···················· | ···················· | ···················· | ···················· | NNNNNNNNNNNNNNNNNNNN | NNNNNNNNNNNNNNNNNNNN | 1 |
| A | 2393 hCoV-19/England/CAMB-7559E/2020\|EPI_ISL_425407\|202 | ···················· | ···················· | ···················· | ···················· | NNNNNNNNNNNNNNNNNNNN | NNNNNNNNNNNNNNNNNNNN | 1 |
| A | 2394 hCoV-19/England/CAMB-755CB/2020\|EPI_ISL_425409\|202 | ···················· | ···················· | ···················· | ···················· | NNNNNNNNNNNNNNNNNNNN | NNNNNNNNNNNNNNNNNNNN | 1 |
| A | 2395 hCoV-19/England/CAMB-755DA/2020\|EPI_ISL_425410\|202 | ···················· | ···················· | ···················· | ···················· | NNNNNNNNNNNNNNNNNNNN | NNNNNNNNNNNNNNNNNNNN | 1 |
| A | 2396 hCoV-19/England/CAMB-755E9/2020\|EPI_ISL_425411\|202 | ···················· | ···················· | ···················· | ···················· | NNNNNNNNNNNNNNNNNNNN | NNNNNNNNNNNNNNNNNNNN | 1 |
| A | 2397 hCoV-19/England/CAMB-755F8/2020\|EPI_ISL_425412\|202 | ···················· | ···················· | ···················· | ···················· | NNNNNNNNNNNNNNNNNNNN | NNNNNNNNNNNNNNNNNNNN | 1 |
| A | 2452 hCoV-19/Australia/VIC409/2020\|EPI_ISL_426711\|2020- | ···················· | ···················· | ···················· | ···················· | NNNNNNNNNNNNNNNNNNNN | ···················· | 1 |
| A | 2453 hCoV-19/Australia/VIC414/2020\|EPI_ISL_426714\|2020- | ···················· | ···················· | ···················· | ···················· | NNNNNNNNNNNNNNNNNNNN | NNNNNNNNNNNNNNNNNNNN | 1 |
| A | 2454 hCoV-19/Australia/VIC415/2020\|EPI_ISL_426715\|2020- | ···················· | ···················· | ···················· | ···················· | ···················· | NNNNNNNNNNNNNNNNNNNN | 1 |
| A | 2455 hCoV-19/Australia/VIC417/2020\|EPI_ISL_426717\|2020- | ···················· | ···················· | ···················· | ···················· | NNNNNNNNNNNNNNNNNNNN | NNNNNNNNNNNNNNNNNNNN | 1 |
| A | 2456 hCoV-19/Australia/VIC418/2020\|EPI_ISL_426718\|2020- | ···················· | ···················· | ···················· | ···················· | NNNNNNNNNNNNNNNNNNNN | NNNNNNNNNNNNNNNNNNNN | 1 |
| A | 2457 hCoV-19/Australia/VIC421/2020\|EPI_ISL_426721\|2020- | ···················· | ···················· | ···················· | ···················· | NNNNNNNNNNNNNNNNNNNN | NNNNNNNNNNNNNNNNNNNN | 1 |
| A | 2458 hCoV-19/Australia/VIC425/2020\|EPI_ISL_426725\|2020- | ···················· | ···················· | ···················· | ···················· | NNNNNNNNNNNNNNNNNNNN | NNNNNNNNNNNNNNNNNNNN | 1 |
| A | 2459 hCoV-19/Australia/VIC427/2020\|EPI_ISL_426727\|2020- | ···················· | ···················· | ···················· | ···················· | NNNNNNNNNNNNNNNNNNNN | NNNNNNNNNNNNNNNNNNNN | 1 |
| A | 2460 hCoV-19/Australia/VIC429/2020\|EPI_ISL_426729\|2020- | ···················· | ···················· | ···················· | ···················· | NNNNNNNNNNNNNNNNNNNN | NNNNNNNNNNNNNNNNNNNN | 1 |
| A | 2461 hCoV-19/Australia/VIC430/2020\|EPI_ISL_426730\|2020- | ···················· | ···················· | ···················· | ···················· | NNNNNNNNNNNNNNNNNNNN | NNNNNNNNNNNNNNNNNNNN | 1 |
| A | 2462 hCoV-19/Australia/VIC433/2020\|EPI_ISL_426732\|2020- | ···················· | ···················· | ···················· | ···················· | NNNNNNNNNNNNNNNNNNNN | NNNNNNNNNNNNNNNNNNNN | 1 |
| A | 2463 hCoV-19/Australia/VIC434/2020\|EPI_ISL_426733\|2020- | ···················· | ···················· | ···················· | ···················· | NNNNNNNNNNNNNNNNNNNN | NNNNNNNNNNNNNNNNNNNN | 1 |
| A | 2464 hCoV-19/Australia/VIC437/2020\|EPI_ISL_426734\|2020- | ···················· | ···················· | ···················· | ···················· | NNNNNNNNNNNNNNNNNNNN | NNNNNNNNNNNNNNNNNNNN | 1 |
| A | 2465 hCoV-19/Australia/VIC438/2020\|EPI_ISL_426735\|2020- | ···················· | ···················· | ···················· | ···················· | NNNNNNNNNNNNNNNNNNNN | NNNNNNNNNNNNNNNNNNNN | 1 |
| A | 2466 hCoV-19/Australia/VIC439/2020\|EPI_ISL_426736\|2020- | ···················· | ···················· | ···················· | ···················· | NNNNNNNNNNNNNNNNNNNN | NNNNNNNNNNNNNNNNNNNN | 1 |
| A | 2467 hCoV-19/Australia/VIC440/2020\|EPI_ISL_426737\|2020- | ···················· | ···················· | ···················· | ···················· | NNNNNNNNNNNNNNNNNNNN | NNNNNNNNNNNNNNNNNNNN | 1 |
| A | 2468 hCoV-19/Australia/VIC443/2020\|EPI_ISL_426740\|2020- | ···················· | ···················· | ···················· | ···················· | NNNNNNNNNNNNNNNNNNNN | NNNNNNNNNNNNNNNNNNNN | 1 |
| A | 2470 hCoV-19/Australia/VIC447/2020\|EPI_ISL_426743\|2020- | ···················· | ···················· | ···················· | ···················· | NNNNNNNNNNNNNNNNNNNN | NNNNNNNNNNNNNNNNNNNN | 1 |
| A | 2472 hCoV-19/Australia/VIC450/2020\|EPI_ISL_426746\|2020- | ···················· | ···················· | ···················· | ···················· | NNNNNNNNNNNNNNNNNNNN | NNNNNNNNNNNNNNNNNNNN | 1 |
| A | 2473 hCoV-19/Australia/VIC451/2020\|EPI_ISL_426747\|2020- | ···················· | ···················· | ···················· | ···················· | NNNNNNNNNNNNNNNNNNNN | NNNNNNNNNNNNNNNNNNNN | 1 |
| A | 2474 hCoV-19/Australia/VIC454/2020\|EPI_ISL_426750\|2020- | ···················· | ···················· | ···················· | ···················· | NNNNNNNNNNNNNNNNNNNN | NNNNNNNNNNNNNNNNNNNN | 1 |
| A | 2475 hCoV-19/Australia/VIC456/2020\|EPI_ISL_426751\|2020- | ···················· | ···················· | ···················· | ···················· | NNNNNNNNNNNNNNNNNNNN | NNNNNNNNNNNNNNNNNNNN | 1 |
| A | 2477 hCoV-19/Australia/VIC460/2020\|EPI_ISL_426753\|2020- | ···················· | ···················· | ···················· | ···················· | NNNNNNNNNNNNNNNNNNNN | NNNNNNNNNNNNNNNNNNNN | 1 |
| A | 2478 hCoV-19/Australia/VIC461/2020\|EPI_ISL_426754\|2020- | ···················· | ···················· | ···················· | ···················· | NNNNNNNNNNNNNNNNNNNN | NNNNNNNNNNNNNNNNNNNN | 1 |
| A | 2479 hCoV-19/Australia/VIC463/2020\|EPI_ISL_426756\|2020- | ···················· | ···················· | ···················· | ···················· | NNNNNNNNNNNNNNNNNNNN | NNNNNNNNNNNNNNNNNNNN | 1 |
| A | 2480 hCoV-19/Australia/VIC468/2020\|EPI_ISL_426761\|2020- | ···················· | ···················· | ···················· | ···················· | NNNNNNNNNNNNNNNNNNNN | NNNNNNNNNNNNNNNNNNNN | 1 |
| A | 2483 hCoV-19/Australia/VIC493/2020\|EPI_ISL_426766\|2020- | ···················· | ···················· | ···················· | ···················· | NNNNNNN············· | ···················· | 1 |
| A | 2485 hCoV-19/Australia/VIC495/2020\|EPI_ISL_426768\|2020- | ···················· | ···················· | ···················· | ···················· | ·NNNNNNNNNNNNNNNNNNN | ···················· | 1 |
| A | 2486 hCoV-19/Australia/VIC496/2020\|EPI_ISL_426769\|2020- | ···················· | ···················· | ···················· | ···················· | NNNNNNN············· | NNNNNNNNNNNNNNNN·NNN | 1 |
| A | 2491 hCoV-19/Australia/VIC503/2020\|EPI_ISL_426774\|2020- | ···················· | ···················· | ···················· | ···················· | NNNNNNNNNNNNNNNNNNNN | ···················· | 1 |
| A | 2729 hCoV-19/England/20140005304/2020\|EPI_ISL_423380\|20 | ···················· | ···················· | ···················· | ···················· | ··N·NN······N··NNN·· | ···················· | 1 |
| A | 2733 hCoV-19/England/20140007604/2020\|EPI_ISL_423384\|20 | ···················· | ···················· | ···················· | ···················· | NNNNNNNNNNNNNNNNNNNN | NNNNNNNNNNNNNNN·N··N | 1 |
| A | 2735 hCoV-19/England/20140009304/2020\|EPI_ISL_423386\|20 | ···················· | ···················· | ···················· | ···················· | NNNNNNNNNNNNNNNNNNNN | NNNNNNNNNNNNNNNNNNNN | 1 |
| A | 2737 hCoV-19/England/20140060304/2020\|EPI_ISL_423389\|20 | ···················· | ···················· | ···················· | ···················· | ··NNN·NNNN·NN·N·NNNN | ···················· | 1 |
| A | 2844 hCoV-19/Iceland/333/2020\|EPI_ISL_417662\|2020-03-14 | ···················· | ···················· | ···················· | ···················· | ·······NNNNNNNNNNNNN | NNNNNNNNNNNNNNNNNNNN | 1 |
| A | 2863 hCoV-19/England/20148003904/2020\|EPI_ISL_422000\|20 | ···················· | ···················· | ···················· | ···················· | NNNN················ | NNNNNN··NNN···N····· | 1 |
| A | 2868 hCoV-19/England/20148026504/2020\|EPI_ISL_422008\|20 | ···················· | ···················· | ···················· | ···················· | ···················· | NNNNNNNNNNNNNNNNNNNN | 1 |
| A | 2870 hCoV-19/England/20148041604/2020\|EPI_ISL_422010\|20 | ···················· | ···················· | ···················· | ···················· | ···················· | NNNN················ | 1 |
| A | 2895 hCoV-19/England/20138063804/2020\|EPI_ISL_423321\|20 | ···················· | ···················· | ···················· | ···················· | ··N·N·N············· | NNNNN··············· | 1 |
| A | 2899 hCoV-19/England/20139015304/2020\|EPI_ISL_423340\|20 | ···················· | ···················· | ···················· | ···················· | ···················· | NNNNNNNNNNNNNNNNNNNN | 1 |
| A | 2900 hCoV-19/England/20139015804/2020\|EPI_ISL_423343\|20 | ···················· | ···················· | ···················· | ···················· | NNNNNNNNNNNNNNNNNNNN | ···········NNNNNNNNN | 1 |
| A | 2902 hCoV-19/England/20139060704/2020\|EPI_ISL_423350\|20 | ···················· | ···················· | ···················· | ···················· | ··NNNNNNNNNNNNNNNNNN | NNNNNNNNNNNNNNN·N··· | 1 |
| A | 2903 hCoV-19/England/20139064404/2020\|EPI_ISL_423351\|20 | ···················· | ···················· | ···················· | ···················· | ···N·······NN······· | NNNNNNNNNNNNNNNNNNNN | 1 |
| A | 2913 hCoV-19/England/20139070204/2020\|EPI_ISL_423361\|20 | ···················· | ···················· | ···················· | ···················· | ·········N····NN···N | ··NNN··············· | 1 |
| A | 2914 hCoV-19/England/20139070504/2020\|EPI_ISL_423362\|20 | ···················· | ···················· | ···················· | ···················· | ·NNNNNNNNNNNNNNNNNNN | N·NNNNNN·NNNNNNNN··· | 1 |
| A | 2915 hCoV-19/England/20139070704/2020\|EPI_ISL_423363\|20 | ···················· | ···················· | ···················· | ···················· | NNNNNNNNNNNNNNNNNNNN | NNNNNNNNNNNNNNNNNNNN | 1 |
| A | 2916 hCoV-19/England/20139071704/2020\|EPI_ISL_423364\|20 | ···················· | ···················· | ···················· | ···················· | ···········N··N····· | ····N·N··N··NN·····N | 1 |

| | | | | | | | | |
|---|---|---|---|---|---|---|---|---|
| A | 2919 hCoV-19/England/20140000404/2020\|EPI_ISL_423367\|20 | ···················· | ···················· | ···················· | ···················· | NNNNNNNNNNNNNNNNNNNN | NNNNNNNNNNNNNNNNNNNN | 1 |
| A | 2921 hCoV-19/England/20140000904/2020\|EPI_ISL_423369\|20 | ···················· | ···················· | ···················· | ···················· | ·N·NNNN·NN·N·······N | NNNNNNNNNNNNNNNNNNNN | 1 |
| A | 2927 hCoV-19/England/20144012404/2020\|EPI_ISL_423507\|20 | ···················· | ···················· | ···················· | ···················· | ··NNNNNNNNNNNNNNNN·· | N······N·······NN··· | 1 |
| A | 2930 hCoV-19/England/20144018904/2020\|EPI_ISL_423512\|20 | ···················· | ···················· | ···················· | ···················· | ···················· | NNNNNNNNNNNNNNNN···· | 1 |
| A | 2931 hCoV-19/England/20144022704/2020\|EPI_ISL_423521\|20 | ···················· | ···················· | ···················· | ···················· | NNNNNNNNNNNNNNNNNNNN | NNNNNNNNNNNNNNNNNNNN | 1 |
| A | 3017 hCoV-19/Scotland/EDB327/2020\|EPI_ISL_425973\|2020-0 | ···················· | ···················· | ···················· | ···················· | ···N····NN····NNN··· | NN·N·····NNN···N··N· | 1 |
| A | 3018 hCoV-19/Scotland/EDB336/2020\|EPI_ISL_425975\|2020-0 | ···················· | ···················· | ···················· | ···················· | NNNNNNNNNNNNNNNNNNNN | NNNNNNNNNNNNNNNNNNNN | 1 |
| A | 3024 hCoV-19/Scotland/EDB367/2020\|EPI_ISL_425984\|2020-0 | ···················· | ···················· | ···················· | ···················· | NNNNNNNNNNNNNNNNNNNN | NNNNNNNNNNNNNNNNNNNN | 1 |
| A | 3041 hCoV-19/Australia/VIC781/2020\|EPI_ISL_427056\|2020- | ···················· | ···················· | ···················· | ···················· | NNNNNNNNNNNNNNNNNNNN | NNNNNNNNNNNNNNNNNNNN | 1 |
| A | 3042 hCoV-19/Australia/VIC782/2020\|EPI_ISL_427057\|2020- | ···················· | ···················· | ···················· | ···················· | NNNNNNNNNNNNNNNNNNNN | NNNNNNNNNNNNNNNNNNNN | 1 |
| A | 3043 hCoV-19/Australia/VIC783/2020\|EPI_ISL_427058\|2020- | ···················· | ···················· | ···················· | ···················· | NNNNNNN············· | ···················· | 1 |
| A | 3045 hCoV-19/Australia/VIC786/2020\|EPI_ISL_427061\|2020- | ···················· | ···················· | ···················· | ···················· | NNNNNNNNNNNNNNNNNNNN | NNNNNNNNNNNNNNNNNNNN | 1 |
| A | 3046 hCoV-19/Australia/VIC787/2020\|EPI_ISL_427062\|2020- | ···················· | ···················· | ···················· | ···················· | NNNNNNNNNNNNNNNNNNNN | NNNNNNNNNNNNNNNNNNNN | 1 |
| A | 3047 hCoV-19/Australia/VIC788/2020\|EPI_ISL_427063\|2020- | ···················· | ···················· | ···················· | ···················· | NNNNNNNNNNNNNNNNNNNN | NNNNNNNNNNNNNNNNNNNN | 1 |
| A | 3048 hCoV-19/Australia/VIC790/2020\|EPI_ISL_427065\|2020- | ···················· | ···················· | ···················· | ···················· | NNNNNNN············· | ···················· | 1 |
| A | 3049 hCoV-19/Australia/VIC791/2020\|EPI_ISL_427066\|2020- | ···················· | ···················· | ···················· | ···················· | NNNNNNNNNNNNNNNNNNNN | NNNNNNNNNNNNNNNNNNNN | 1 |
| A | 3050 hCoV-19/Australia/VIC793/2020\|EPI_ISL_427068\|2020- | ···················· | ···················· | ···················· | ···················· | ··NNNNNNNNNNNNNNNNNN | NNNNNNNNNNNNNNNNNNNN | 1 |
| A | 3051 hCoV-19/Australia/VIC794/2020\|EPI_ISL_427069\|2020- | ···················· | ···················· | ···················· | ···················· | NNNNNNNNNNNNNNNNNNNN | NNNNNNNNNNNNNNNNNNNN | 1 |
| A | 3052 hCoV-19/Australia/VIC796/2020\|EPI_ISL_427070\|2020- | ···················· | ···················· | ···················· | ···················· | ···················· | NNNN················ | 1 |
| A | 3053 hCoV-19/Australia/VIC797/2020\|EPI_ISL_427071\|2020- | ···················· | ···················· | ···················· | ···················· | NNNNNNN············· | NNNN················ | 1 |
| A | 3054 hCoV-19/Australia/VIC798/2020\|EPI_ISL_427072\|2020- | ···················· | ···················· | ···················· | ···················· | ···················· | NNNNNNNNNNNNNNNNNNNN | 1 |
| A | 3055 hCoV-19/Australia/VIC799/2020\|EPI_ISL_427073\|2020- | ···················· | ···················· | ···················· | ···················· | NNNNNNNNNNNNNNNNNNNN | NNNNNNNNN·····NNNNNN | 1 |
| A | 3056 hCoV-19/Australia/VIC800/2020\|EPI_ISL_427074\|2020- | ···················· | ···················· | ···················· | ···················· | ···················· | NNNN················ | 1 |
| A | 3057 hCoV-19/Australia/VIC802/2020\|EPI_ISL_427076\|2020- | ···················· | ···················· | ···················· | ···················· | NNNNNNNNNNNNNNNNNNNN | NNNNNNNNNNNNNNNNNNNN | 1 |
| A | 3058 hCoV-19/Australia/VIC804/2020\|EPI_ISL_427078\|2020- | ···················· | ···················· | ···················· | ···················· | NNNNNNNNNNNNNNNNNNNN | NNNNNNNNNNNNNNNNNNNN | 1 |
| A | 3059 hCoV-19/Australia/VIC805/2020\|EPI_ISL_427079\|2020- | ···················· | ···················· | ···················· | ···················· | NNNNNNNNNNNNNNNNNNNN | NNNNNNNNNNNNNNNNNNNN | 1 |
| A | 3060 hCoV-19/Australia/VIC806/2020\|EPI_ISL_427080\|2020- | ···················· | ···················· | ···················· | ···················· | ···················· | NNNN················ | 1 |
| A | 3097 hCoV-19/Shanghai/SH0041/2020\|EPI_ISL_416350\|2020-0 | ···················· | ···················· | ···················· | ···················· | ···················· | ··NNNNN············· | 1 |
| A | 3139 hCoV-19/Iceland/328/2020\|EPI_ISL_417657\|2020-03-13 | ···················· | ···················· | ···················· | ···················· | ······NNNNNNNNNNNNNN | NNNNNNNNNNNNNNNNNNNN | 1 |
| A | 3141 hCoV-19/Iceland/331/2020\|EPI_ISL_417660\|2020-03-14 | ···················· | ···················· | ···················· | ···················· | ···················· | NN·····N·······NNNN· | 1 |
| A | 3149 hCoV-19/Iceland/88/2020\|EPI_ISL_417675\|2020-03-10 | ···················· | ···················· | ···················· | ···················· | ···················· | NNNNNNNNNNNNNNNNNNNN | 1 |
| A | 3153 hCoV-19/Iceland/79/2020\|EPI_ISL_417679\|2020-03-09 | ···················· | ···················· | ···················· | ···················· | ···················· | NNNNNNNNNNNNNNNNNNNN | 1 |
| A | 3170 hCoV-19/Iceland/35/2020\|EPI_ISL_417697\|2020-03-04 | ···················· | ···················· | ···················· | ···················· | ···················· | ·NNN···NN··········· | 1 |
| A | 3193 hCoV-19/Australia/VIC716/2020\|EPI_ISL_427001\|2020- | ···················· | ···················· | ···················· | ···················· | ···················· | NNNNNNNNNNNNNNNNNNNN | 1 |
| A | 3195 hCoV-19/Australia/VIC720/2020\|EPI_ISL_427003\|2020- | ···················· | ···················· | ···················· | ···················· | NNNNNNNNNNNNNNNNNNNN | NNNNNNNNNNNNNNNNNNNN | 1 |
| A | 3196 hCoV-19/Australia/VIC725/2020\|EPI_ISL_427006\|2020- | ···················· | ···················· | ···················· | ···················· | NNNNNNNNNNNNNNNNNNNN | NNNNNNNNNNNNNNNNNNNN | 1 |
| A | 3197 hCoV-19/Australia/VIC727/2020\|EPI_ISL_427007\|2020- | ···················· | ···················· | ···················· | ···················· | NNNNNNNNNNNNNNNNNNNN | NNNNNNNNNNNNNNNNNNNN | 1 |
| A | 3199 hCoV-19/Australia/VIC732/2020\|EPI_ISL_427011\|2020- | ···················· | ···················· | ···················· | ···················· | NNNNNNNNNNNNNNNNNNNN | ···················· | 1 |
| A | 3200 hCoV-19/Australia/VIC733/2020\|EPI_ISL_427012\|2020- | ···················· | ···················· | ···················· | ···················· | NNNNNNN············· | ···················· | 1 |
| A | 3202 hCoV-19/Australia/VIC735/2020\|EPI_ISL_427014\|2020- | ···················· | ···················· | ···················· | ···················· | NNNNNNNNNNNNNNNNNNNN | NNNNNNNNNNNNNNNNNNNN | 1 |
| A | 3203 hCoV-19/Australia/VIC736/2020\|EPI_ISL_427015\|2020- | ···················· | ···················· | ···················· | ···················· | NNNNNNNNNNNNNNNNNNNN | NNNNNNNNNNNNNNNNNNNN | 1 |
| A | 3204 hCoV-19/Australia/VIC737/2020\|EPI_ISL_427016\|2020- | ···················· | ···················· | ···················· | ···················· | ··················NN | NNNNNNNNNNNNNNNNNNNN | 1 |
| A | 3205 hCoV-19/Australia/VIC738/2020\|EPI_ISL_427017\|2020- | ···················· | ···················· | ···················· | ···················· | ···················· | NNNNNNNNNNNNNNNNNNNN | 1 |
| A | 3206 hCoV-19/Australia/VIC739/2020\|EPI_ISL_427018\|2020- | ···················· | ···················· | ···················· | ···················· | NNNNNNNNNNNNNNNNNNNN | NNNNNNNNNNNNNNNNNNNN | 1 |
| A | 3208 hCoV-19/Australia/VIC742/2020\|EPI_ISL_427020\|2020- | ···················· | ···················· | ···················· | ···················· | NNNNNNNNNNNNNNNNNNNN | NNNN················ | 1 |
| A | 3209 hCoV-19/Australia/VIC744/2020\|EPI_ISL_427022\|2020- | ···················· | ···················· | ···················· | ···················· | ···················· | NNNN················ | 1 |
| A | 3210 hCoV-19/Australia/VIC745/2020\|EPI_ISL_427023\|2020- | ···················· | ···················· | ···················· | ···················· | NNNNNNNNNNNNNNNNNNNN | ···················· | 1 |
| A | 3211 hCoV-19/Australia/VIC746/2020\|EPI_ISL_427024\|2020- | ···················· | ···················· | ···················· | ···················· | NNNNNNNNNNNNNNNNNNNN | NNNNNNNNN········NNN | 1 |
| A | 3212 hCoV-19/Australia/VIC747/2020\|EPI_ISL_427025\|2020- | ···················· | ···················· | ···················· | ···················· | ···················· | NNNNNNNNNNNNNNNNNNNN | 1 |
| A | 3213 hCoV-19/Australia/VIC748/2020\|EPI_ISL_427026\|2020- | ···················· | ···················· | ···················· | ···················· | NNNNNNNNNNNNNNNNNNNN | NNNNNNNNNNNNNNNNNNNN | 1 |
| A | 3214 hCoV-19/Australia/VIC751/2020\|EPI_ISL_427028\|2020- | ···················· | ···················· | ···················· | ···················· | ·NNNNNN············· | NNNNN··············· | 1 |
| A | 3216 hCoV-19/Australia/VIC754/2020\|EPI_ISL_427031\|2020- | ···················· | ···················· | ···················· | ···················· | NNNNNNNNNNNNNNNNNNNN | NNNNNNNNNNNNNNNNNNNN | 1 |
| A | 3217 hCoV-19/Australia/VIC760/2020\|EPI_ISL_427036\|2020- | ···················· | ···················· | ···················· | ···················· | NNNNNNNNNNNNNNNNNNNN | NNNNNNNNNNNNNNNNNNNN | 1 |
| A | 3218 hCoV-19/Australia/VIC762/2020\|EPI_ISL_427038\|2020- | ···················· | ···················· | ···················· | ···················· | ···················· | NNNNNNNNNNNNNNNNNNNN | 1 |
| A | 3219 hCoV-19/Australia/VIC764/2020\|EPI_ISL_427040\|2020- | ···················· | ···················· | ···················· | ···················· | NNNNNNNNNNNNNNNNNNNN | NNNNNNNNNNNNNNNNNNNN | 1 |
| A | 3220 hCoV-19/Australia/VIC765/2020\|EPI_ISL_427041\|2020- | ···················· | ···················· | ···················· | ···················· | NNNNNNNNNNNNNNNNNNNN | NNNNNNNNNNNNNNNN···· | 1 |
| A | 3222 hCoV-19/Australia/VIC768/2020\|EPI_ISL_427045\|2020- | ···················· | ···················· | ···················· | ···················· | NNNNNNN············· | NNNN················ | 1 |
| A | 3223 hCoV-19/Australia/VIC769/2020\|EPI_ISL_427046\|2020- | ···················· | ···················· | ···················· | ···················· | ···················· | NNNNNNNNNNNNNNNNNNNN | 1 |
| A | 3224 hCoV-19/Australia/VIC772/2020\|EPI_ISL_427048\|2020- | ···················· | ···················· | ···················· | ···················· | NNNNNNNNNNNNNNNNNNNN | NNNNNNNNN··········· | 1 |
| A | 3225 hCoV-19/Australia/VIC773/2020\|EPI_ISL_427049\|2020- | ···················· | ···················· | ···················· | ···················· | NNNNNNNNNNNNNNNNNNNN | NNNNNNNNN··········· | 1 |
| A | 3505 hCoV-19/England/20140069104/2020\|EPI_ISL_423403\|20 | ···················· | ···················· | ···················· | ···················· | ······NNNNNNNNNNNN·· | ···················· | 1 |
| A | 3507 hCoV-19/England/20140070004/2020\|EPI_ISL_423405\|20 | ···················· | ···················· | ···················· | ···················· | NNNNNNNNNNNNNNNNNNNN | NNNNNNNNNNNNNNNNNNNN | 1 |
| A | 3508 hCoV-19/England/20140070304/2020\|EPI_ISL_423406\|20 | ···················· | ···················· | ···················· | ···················· | NNNNNNNNNNNNNNNNNNNN | NNNNNNNNNNNNNNNNNNNN | 1 |
| A | 3513 hCoV-19/England/20140071404/2020\|EPI_ISL_423412\|20 | ···················· | ···················· | ···················· | ···················· | ·NN····NNNNN········ | ···················· | 1 |
| A | 3519 hCoV-19/England/20140073904/2020\|EPI_ISL_423419\|20 | ···················· | ···················· | ···················· | ···················· | ··NNNNNNNNNNNNNNNNNN | NNNNNNNNNNNNNNNN···N | 1 |
| A | 3520 hCoV-19/England/20140074104/2020\|EPI_ISL_423420\|20 | ···················· | ···················· | ···················· | ···················· | NNNNN·N············· | NNNNNNNNNNNNNNNNNNNN | 1 |
| A | 3521 hCoV-19/England/20142002004/2020\|EPI_ISL_423421\|20 | ···················· | ···················· | ···················· | ···················· | ··N················· | NNNNNN·N··NN····N··· | 1 |
| A | 3522 hCoV-19/England/20142002104/2020\|EPI_ISL_423422\|20 | ···················· | ···················· | ···················· | ···················· | NNNNNNNNNNNNNNNNNNNN | ···················· | 1 |
| A | 3523 hCoV-19/England/20142002204/2020\|EPI_ISL_423423\|20 | ···················· | ···················· | ···················· | ···················· | NNNNNNNNNNNNNNNNNNNN | NNNNNNNNNNNNNNNNNNNN | 1 |
| A | 3527 hCoV-19/England/20142012304/2020\|EPI_ISL_423428\|20 | ···················· | ···················· | ···················· | ···················· | NNNNNNNNNNNNNNNNNNNN | NNNNNNNNNNNNNNNNNNNN | 1 |
| A | 3534 hCoV-19/England/20142018604/2020\|EPI_ISL_423440\|20 | ···················· | ···················· | ···················· | ···················· | ··NNNNNNNNNNNNNNNNNN | ···················· | 1 |
| A | 3536 hCoV-19/England/20142019004/2020\|EPI_ISL_423442\|20 | ···················· | ···················· | ···················· | ···················· | ··N·····N·····N·N··N | ····N··············· | 1 |
| A | 3537 hCoV-19/England/20142019504/2020\|EPI_ISL_423443\|20 | ···················· | ···················· | ···················· | ···················· | ············N·NNNNN· | NNNNNNN············· | 1 |
| A | 3540 hCoV-19/England/20142053804/2020\|EPI_ISL_423458\|20 | ···················· | ···················· | ···················· | ···················· | NNNNNNNNNNNNNNNNNNNN | NNNNNNNNNNNNNNNNNNNN | 1 |
| A | 3541 hCoV-19/England/20142078304/2020\|EPI_ISL_423459\|20 | ···················· | ···················· | ···················· | ···················· | NNNNNNNNNNN····N··NN | NNNNNNNNNNNNNNNNNNNN | 1 |
| A | 3544 hCoV-19/England/20142082704/2020\|EPI_ISL_423463\|20 | ···················· | ···················· | ···················· | ···················· | ···················· | NNNNNNNNNNNNNNNN···· | 1 |
| A | 3549 hCoV-19/England/20144005404/2020\|EPI_ISL_423490\|20 | ···················· | ···················· | ···················· | ···················· | NNNNNNNNNNNNNNNNNNNN | NNNNNNNNNNNNNNNNNNNN | 1 |
| A | 3550 hCoV-19/England/20144005904/2020\|EPI_ISL_423491\|20 | ···················· | ···················· | ···················· | ···················· | ···················· | NNNNNNNNNNNNNNNNNNK· | 1 |
| A | 3551 hCoV-19/England/20144006004/2020\|EPI_ISL_423492\|20 | ···················· | ···················· | ···················· | ···················· | NNNNNNNNNNNNNNNNNNNN | NNNNNNNNNNNNNNNNNNNN | 1 |
| A | 3553 hCoV-19/England/20144006404/2020\|EPI_ISL_423494\|20 | ···················· | ···················· | ···················· | ···················· | NNNNNNNNNNNNNNNNNNNN | NNNN················ | 1 |

| | | | | | | | | | |
|---|---|---|---|---|---|---|---|---|---|
| A | 3558 | hCoV-19/England/20144008204/2020\|EPI_ISL_423499\|20 | ···················· | ···················· | ···················· | ···················· | ···················· | NNNNNNNNNNNNNNNNNNNN | 1 |
| A | 3596 | hCoV-19/England/20112112106/2020\|EPI_ISL_417314\|20 | ···················· | ···················· | ···················· | ···················· | ··NNNN·NNN·NN···N··N | NNNNNNNNNNNNNNN····· | 1 |
| A | 3639 | hCoV-19/Belgium/ULG-7134/2020\|EPI_ISL_418628\|2020- | ···················· | ···················· | ···················· | ···················· | ··············NN···· | NN·N···NN··········· | 1 |
| A | 3644 | hCoV-19/Belgium/ULG-7626/2020\|EPI_ISL_418633\|2020- | ···················· | ···················· | ···················· | ···················· | ········N····NN····· | NN·N···NN·······N··· | 1 |
| A | 3654 | hCoV-19/Belgium/ULG-9619/2020\|EPI_ISL_418649\|2020- | ···················· | ···················· | ···················· | ···················· | ···N··········N····· | ···N···NN·N··NN····· | 1 |
| A | 3754 | hCoV-19/England/20142023804/2020\|EPI_ISL_423449\|20 | ···················· | ···················· | ···················· | ···················· | ···················· | NNNNNNNNNNNNNNNNNNNN | 1 |
| A | 3755 | hCoV-19/England/20142023904/2020\|EPI_ISL_423450\|20 | ···················· | ···················· | ···················· | ···················· | ··NN····NNN·······NN | ···N············NNNN | 1 |
| A | 3756 | hCoV-19/England/20142024404/2020\|EPI_ISL_423451\|20 | ···················· | ···················· | ···················· | ···················· | ·NN·····N····NN····· | ···················· | 1 |
| A | 3757 | hCoV-19/England/20142024704/2020\|EPI_ISL_423452\|20 | ···················· | ···················· | ···················· | ···················· | ·NNNNNNNNNNNNNNNNNNN | NNNNNNNNNNNNNNNNNNNN | 1 |
| A | 3759 | hCoV-19/England/20142026804/2020\|EPI_ISL_423455\|20 | ···················· | ···················· | ···················· | ···················· | NNNNNNNNNNNNNNNNNNNN | NNNNNNNNNNNNNNNNNNNN | 1 |
| A | 3760 | hCoV-19/England/20142053204/2020\|EPI_ISL_423457\|20 | ···················· | ···················· | ···················· | ···················· | NNNNNNNNNNNNNNNNNNNN | NNNNNNNNNNNNNNNNNNNN | 1 |
| A | 3761 | hCoV-19/England/20142131504/2020\|EPI_ISL_423469\|20 | ········T··········· | ···················· | ···················· | ···················· | NNNNNNNNNNNNNNNNNNNN | ···················· | 1 |
| A | 3762 | hCoV-19/England/20142131704/2020\|EPI_ISL_423471\|20 | ···T················ | ···················· | ···················· | ···················· | ····NNNNNNNNNNNNNNNN | NNNNNNNNNNNNNNNNNNNN | 1 |
| A | 3765 | hCoV-19/England/20144000804/2020\|EPI_ISL_423474\|20 | ···················· | ···················· | ···················· | ···················· | NNNNNNNNNNNNNNNNNNNN | NNNNNNNNNNNNNNNNNNNN | 1 |
| A | 3767 | hCoV-19/England/20144001404/2020\|EPI_ISL_423477\|20 | ···················· | ···················· | ···················· | ···················· | ··N·NNNNNNNNNNNNNNNN | NNNNNNNNNNNNNNNNNNNN | 1 |
| A | 3769 | hCoV-19/England/20144003404/2020\|EPI_ISL_423480\|20 | ···················· | ···················· | ···················· | ···················· | NNNNNNNNNNNNNNNNNNNN | NNNNNNNNNNNNNNNNNNNN | 1 |
| A | 3771 | hCoV-19/England/20144003804/2020\|EPI_ISL_423483\|20 | ···················· | ···················· | ···················· | ···················· | ··NNNNNNNNNNN······· | ···········NNNN····· | 1 |
| A | 3794 | hCoV-19/Scotland/EDB014/2020\|EPI_ISL_425820\|2020-0 | ···················· | ···················· | ···················· | ···················· | ···N··········N····· | NN·N·N·NNN···NN·N·N· | 1 |
| A | 3852 | hCoV-19/France/ARA-06042/2020\|EPI_ISL_417333\|2020- | ···················· | ···················· | ···················· | NNNNNNNNNNNNNNNNNNNN | ···················· | ···················· | 1 |
| A | 3897 | hCoV-19/England/20124001302/2020\|EPI_ISL_418684\|20 | ···················· | ···················· | ···················· | ···················· | NNNNNNNNNNNNNNNNNNNN | NNNNNNNNNNNNNNNNNNNN | 1 |
| A | 3898 | hCoV-19/England/20124001702/2020\|EPI_ISL_418685\|20 | ···················· | ···················· | ···················· | ···················· | NNNNNNNNNNNNNNNNNNNN | NNNNNNNNNNNNNNNNNNNN | 1 |
| A | 3899 | hCoV-19/England/20124001902/2020\|EPI_ISL_418686\|20 | ···················· | ···················· | ···················· | ···················· | ·······NNNNNNNNNNNNN | NNNNNNNNNNNNNNNNNNNN | 1 |
| A | 3901 | hCoV-19/England/20124002102/2020\|EPI_ISL_418688\|20 | ···················· | ···················· | ···················· | ···················· | ······NNNNNNNNNNNNNN | ···NN··············· | 1 |
| A | 3905 | hCoV-19/England/20124008902/2020\|EPI_ISL_418694\|20 | ···················· | ···················· | ···················· | ···················· | ··NN·N····N·······N· | ··N·N··············· | 1 |
| A | 3907 | hCoV-19/England/20124011302/2020\|EPI_ISL_418697\|20 | ···················· | ···················· | ···················· | ···················· | ·····NNNNNNNNNNNNNNN | NNNNNNNNNNNNNNNNNNNN | 1 |
| A | 3916 | hCoV-19/Australia/VIC212/2020\|EPI_ISL_419907\|2020- | ···················· | ···················· | ···················· | ···················· | NNNNNNNNNNNNNNNNNNNN | NNNNNNNNNNNNNNNNNNNN | 1 |
| A | 3961 | hCoV-19/Germany/BY-MVP-V2010837/2020\|EPI_ISL_42090 | ···················· | ···················· | ···················· | ···················· | NNNNNNNNNNNNNNNNNNNN | NNNNNNNNNNNNNNNNNNNN | 1 |
| A | 3965 | hCoV-19/Germany/BY-MVP-V2011672/2020\|EPI_ISL_42090 | ···················· | ···················· | ···················· | ···················· | ·NN···········N····· | NN·N·········N······ | 1 |
| A | 3980 | hCoV-19/England/20134025504/2020\|EPI_ISL_423130\|20 | ···················· | ···················· | ···················· | ···················· | NNNNNNNNNNNNNNNNNNNN | NNNNNNNNNNNNNNNNNNNN | 1 |
| A | 3983 | hCoV-19/England/20134026904/2020\|EPI_ISL_423134\|20 | ···················· | ···················· | ···················· | ···················· | ·NN··NNNNNNNNNNNNNNN | ···················· | 1 |
| A | 3987 | hCoV-19/England/20134047404/2020\|EPI_ISL_423142\|20 | ···················· | ···················· | ···················· | ···················· | ·NNNNNNNNNNNNN······ | ·········N·········· | 1 |
| A | 3988 | hCoV-19/England/20134059604/2020\|EPI_ISL_423144\|20 | ···················· | ···················· | ···················· | ···················· | ··NNNNNNNNNNNNNNNNNN | NNNNNNNNNNNNNNNNNNNN | 1 |
| A | 3989 | hCoV-19/England/20134059704/2020\|EPI_ISL_423145\|20 | ···················· | ···················· | ···················· | ···················· | NNNNNNNNNNNNNNNNNNNN | NNNNNNNNNNNNNNNNNNNN | 1 |
| A | 3991 | hCoV-19/England/20134060804/2020\|EPI_ISL_423148\|20 | ···················· | ···················· | ···················· | ···················· | ·NNNNNNNNN·········· | ···················· | 1 |
| A | 3997 | hCoV-19/England/20134067904/2020\|EPI_ISL_423156\|20 | ···················· | ···················· | ···················· | ···················· | ··NNNN···NN·N·N·N·NN | ···················· | 1 |
| A | 4000 | hCoV-19/England/20134068704/2020\|EPI_ISL_423159\|20 | ···················· | ···················· | ···················· | ···················· | NNNNNNNNNNNNNNNNNNNN | NNNNNNNNNNNNNNNNNNNN | 1 |
| A | 4002 | hCoV-19/England/20134069004/2020\|EPI_ISL_423161\|20 | ···················· | ···G················ | ···················· | ···················· | ·NNNNNN·NNNN·NNNNN·N | N·N·NNNN···NN·N·N·NN | 1 |
| A | 4004 | hCoV-19/England/20134072104/2020\|EPI_ISL_423164\|20 | ···················· | ···················· | ···················· | ···················· | NNNNNNNNNNNNNNNNNNNN | NNNNNNNNNNNNNNNNNNNN | 1 |
| A | 4005 | hCoV-19/England/20134073904/2020\|EPI_ISL_423168\|20 | ···················· | ···················· | ···················· | ···················· | ··NN···NNNNNNNNNNNNN | NNNNNNNNNNNNNNNN···N | 1 |
| A | 4008 | hCoV-19/England/20134078204/2020\|EPI_ISL_423173\|20 | ···················· | ···················· | ···················· | ···················· | NNNNNNNNNNNNNNNNNNNN | NNNNNNNNNNNNNNNNNNNN | 1 |
| A | 4010 | hCoV-19/England/20134078604/2020\|EPI_ISL_423175\|20 | ···················· | ···················· | ···················· | ···················· | ·NNNNNNNNNNNNNNNNNNN | ···················· | 1 |
| A | 4012 | hCoV-19/England/20134078904/2020\|EPI_ISL_423177\|20 | ···················· | ···················· | ···················· | ···················· | ·NNNNNNNNNNNNNNNNNNN | NNNNNNNNNNNNNNNNNNNN | 1 |
| A | 4016 | hCoV-19/England/201340875/2020\|EPI_ISL_423193\|2020 | ···················· | ···················· | ···················· | ···················· | ···················· | ··NNN··NN····N······ | 1 |
| A | 4021 | hCoV-19/England/201360037/2020\|EPI_ISL_423199\|2020 | ···················· | ···················· | ···················· | ···················· | ···················· | NNNNN···NNNNNNNN·NNN | 1 |
| A | 4133 | hCoV-19/England/20134019904/2020\|EPI_ISL_423107\|20 | ···················· | ···················· | ···················· | ···················· | ··NNNNNNNNNNNN··N··N | NNNNNNNNNNNNNNN·NN·N | 1 |
| A | 4137 | hCoV-19/England/20134020904/2020\|EPI_ISL_423111\|20 | ···················· | ·A·················· | ···················· | ···················· | ··NNN··············· | NNNNNNNNNNNN··N····· | 1 |
| A | 4144 | hCoV-19/England/20134023004/2020\|EPI_ISL_423121\|20 | ···················· | ···················· | ···················· | ···················· | ··NNNNN·NNNNNNN·NN·· | ··N···N············· | 1 |
| A | 4147 | hCoV-19/England/20134023704/2020\|EPI_ISL_423124\|20 | ···················· | ···················· | ···················· | ···················· | ·NNNNNNNNNNNNNNNNNNN | NNNNNNNNNN·NNNN··N·· | 1 |
| A | 4295 | hCoV-19/Scotland/EDB114/2020\|EPI_ISL_425901\|2020-0 | ···················· | ···················· | ···················· | ···················· | NNNNNNNNNNNNNNNNNNNN | NNNNNNNNNNNNNNNNNNNN | 1 |
| A | 4297 | hCoV-19/Scotland/EDB119/2020\|EPI_ISL_425903\|2020-0 | ···················· | ···················· | ···················· | ···················· | NNNNNNNNNNNNNNNNNNNN | NNNNNNNNNNNNNNNNNNNN | 1 |
| A | 4299 | hCoV-19/Scotland/EDB122/2020\|EPI_ISL_425905\|2020-0 | ···················· | ···················· | ···················· | ···················· | NNNNNNNNNNNNNNNNNNNN | NNNNNNNNNNNNNNNNNNNN | 1 |
| A | 4302 | hCoV-19/Scotland/EDB128/2020\|EPI_ISL_425908\|2020-0 | ···················· | ···················· | ···················· | ···················· | NNNNNNNNNNNNNNNNNNNN | NNNNNNNNNNNNNNNNNNNN | 1 |
| A | 4305 | hCoV-19/Scotland/EDB144/2020\|EPI_ISL_425913\|2020-0 | ···················· | ···················· | ···················· | ···················· | ···N··········NN···· | NN·····N··NN·NN····· | 1 |
| A | 4313 | hCoV-19/Scotland/EDB183/2020\|EPI_ISL_425929\|2020-0 | ···················· | ···················· | ···················· | ···················· | NNNNNNNNNNNNNNNNNNNN | NNNNNNNNNNNNNNNNNNNN | 1 |
| A | 4314 | hCoV-19/Scotland/EDB185/2020\|EPI_ISL_425930\|2020-0 | ···················· | ···················· | ···················· | ···················· | NNNNNNNNNNNNNNNNNNNN | NNNNNNNNNNNNNNNNNNNN | 1 |
| A | 4315 | hCoV-19/Scotland/EDB186/2020\|EPI_ISL_425931\|2020-0 | ···················· | ···················· | ···················· | ···················· | NNNNNNNNNNNNNNNNNNNN | NNNNNNNNNNNNNNNNNNNN | 1 |
| A | 4318 | hCoV-19/Scotland/EDB191/2020\|EPI_ISL_425934\|2020-0 | ···················· | ···················· | ···················· | ···················· | NNNNNNNNNNNNNNNNNNNN | NNNNNNNNNNNNNNNNNNNN | 1 |
| A | 4319 | hCoV-19/Scotland/EDB192/2020\|EPI_ISL_425935\|2020-0 | ···················· | ···················· | ···················· | ···················· | NNNNNNNNNNNNNNNNNNNN | NNNNNNNNNNNNNNNNNNNN | 1 |
| A | 4321 | hCoV-19/Scotland/EDB194/2020\|EPI_ISL_425937\|2020-0 | ···················· | ···················· | ···················· | ···················· | N·············N····· | ···N···NN·····N····· | 1 |
| A | 4322 | hCoV-19/Scotland/EDB197/2020\|EPI_ISL_425938\|2020-0 | ···················· | ···················· | ···················· | ···················· | NNNNNNNNNNNNNNNNNNNN | NNNNNNNNNNNNNNNNNNNN | 1 |
| A | 4324 | hCoV-19/Scotland/EDB199/2020\|EPI_ISL_425940\|2020-0 | ···················· | ·············A······ | ···················· | ···················· | NNNNNNNNNNNNNNNNNNNN | NNNNNNNNNNNNNNNNNNNN | 1 |
| A | 4325 | hCoV-19/Scotland/EDB202/2020\|EPI_ISL_425941\|2020-0 | ···················· | ···················· | ···················· | ···················· | NNNNNNNNNNNNNNNNNNNN | NNNNNNNNNNNNNNNNNNNN | 1 |
| A | 4326 | hCoV-19/Scotland/EDB204/2020\|EPI_ISL_425942\|2020-0 | ···················· | ···················· | ···················· | ···················· | ·······NN····NNNN·NN | NN·N···NN·NN·······N | 1 |
| A | 4400 | hCoV-19/DRC/KN-0017/2020\|EPI_ISL_417433\|2020-03-11 | ···················· | ···················· | ···················· | ···················· | NNNNNNNNNNNNNNNNNNNN | NNNNNNNNNNNNNNNNNNNN | 1 |
| A | 4402 | hCoV-19/DRC/KN-0043/2020\|EPI_ISL_417435\|2020-03-15 | ···················· | ···················· | ···················· | ···················· | NNN··N··N·····NNNNNN | NNNN···NNNN··NN·N·N· | 1 |
| A | 4463 | hCoV-19/England/20124035802/2020\|EPI_ISL_418715\|20 | ···················· | ···················· | ···················· | ···················· | ···················· | NNNNNNNNNNNNNNNNNNNN | 1 |
| A | 4464 | hCoV-19/England/20124036002/2020\|EPI_ISL_418716\|20 | ···················· | ···················· | ···················· | ···················· | ···················· | ·NN·N···N··········· | 1 |
| A | 4466 | hCoV-19/England/20124037302/2020\|EPI_ISL_418720\|20 | ···················· | ···················· | ···················· | ···················· | ·NNNNNNNNNNNNNNNNNNN | NNNNNNNNNNNNNNNNNNNN | 1 |
| A | 4467 | hCoV-19/England/20124048202/2020\|EPI_ISL_418722\|20 | ···················· | ···················· | ···················· | ···················· | ·NNNNNNNNNNNNNNNNNNN | NNNNNNNNNNN···NNN··· | 1 |
| A | 4470 | hCoV-19/England/20124096802/2020\|EPI_ISL_418733\|20 | ···················· | ···················· | ···················· | ···················· | ···········N········ | ··N·N····NNNN··N···· | 1 |
| A | 4472 | hCoV-19/England/20124099602/2020\|EPI_ISL_418736\|20 | ···················· | ···················· | ···················· | ···················· | NNNNNNNNNNNNNNNNNNNN | NNNNNNNNNNNNNNNNNNNN | 1 |
| A | 4473 | hCoV-19/England/20124100002/2020\|EPI_ISL_418737\|20 | ···················· | ···················· | ···················· | ···················· | ··NNNN·NNN·NN······· | ···················· | 1 |
| A | 4475 | hCoV-19/England/20126006702/2020\|EPI_ISL_418748\|20 | ···················· | ···················· | ···················· | ···················· | ·NNNNNNNNNNNNNNNNNNN | NNNNNNNNNNNNNNNNNNNN | 1 |
| A | 4476 | hCoV-19/England/20126006802/2020\|EPI_ISL_418749\|20 | ···················· | ···················· | ···················· | ···················· | ·······NN········NNN | ···················· | 1 |
| A | 4515 | hCoV-19/England/201360229/2020\|EPI_ISL_423255\|2020 | ···················· | ···················· | ···················· | ···················· | ···················· | NNNN·······N·NN····· | 1 |
| A | 4517 | hCoV-19/England/201360249/2020\|EPI_ISL_423258\|2020 | ···················· | ···················· | ···················· | ···················· | ·NNNNNNNNNNNNNNNNNNN | NNNNNNNNNNNNNNNNNNNN | 1 |
| A | 4525 | hCoV-19/England/201360538/2020\|EPI_ISL_423266\|2020 | ···················· | ···················· | ···················· | ···················· | ···················· | ·············NNNNNNN | 1 |
| A | 4527 | hCoV-19/England/20138009704/2020\|EPI_ISL_423275\|20 | ···················· | ···················· | ···················· | ···················· | NNNNNNNNNNNNNNNNNNNN | NNNNNNNNNNNNNNNNNNNN | 1 |
| A | 4560 | hCoV-19/Scotland/EDB028/2020\|EPI_ISL_425834\|2020-0 | ···················· | ···················· | ···················· | ···················· | NNNNNNNNNNNNNNNNNNNN | NNNNNNNNNNNNNNNNNNNN | 1 |
| A | 4567 | hCoV-19/Scotland/EDB161/2020\|EPI_ISL_425923\|2020-0 | ···················· | ···················· | ···················· | ···················· | NNNNNNNNNNNNNNNNNNNN | NNNNNNNNNNNNNNNNNNNN | 1 |

| | | | | | | | | | |
|---|---|---|---|---|---|---|---|---|---|
| A | 4570 | hCoV-19/Scotland/EDB171/2020\|EPI_ISL_425926\|2020-0 | ···················· | ···················· | ···················· | ···················· | NNNNNNNNNNNNNNNNNNNN | NNNNNNNNNNNNNNNNNNNN | 1 |
| A | 4572 | hCoV-19/Scotland/EDB182/2020\|EPI_ISL_425928\|2020-0 | ···················· | ···················· | ···················· | ···················· | NNNNNNNNNNNNNNNNNNNN | NNNNNNNNNNNNNNNNNNNN | 1 |
| A | 4664 | hCoV-19/England/201360052/2020\|EPI_ISL_423202\|2020 | ···················· | ···················· | ···················· | ···················· | ··NNNNNNNNNNNNNNNNNN | NNNNNNNNNNNNNNNNNNNN | 1 |
| A | 4667 | hCoV-19/England/201360057/2020\|EPI_ISL_423205\|2020 | ···················· | ···················· | ···················· | ···················· | ·NNNNNNNNNNNNNNNNNNN | NNNNNNNNNNNNNNNNNNNN | 1 |
| A | 4668 | hCoV-19/England/201360069/2020\|EPI_ISL_423208\|2020 | ···················· | ···················· | ···················· | ···················· | ···NN··N·········NNN | NNNNNNNNNNNNNNNNNNNN | 1 |
| A | 4669 | hCoV-19/England/201360071/2020\|EPI_ISL_423209\|2020 | ···················· | ···················· | ···················· | ···················· | ·NNNNNNNNNNNNNNNNNNN | ···················· | 1 |
| A | 4683 | hCoV-19/England/201360188/2020\|EPI_ISL_423233\|2020 | ···················· | ···················· | ···················· | ···················· | NNNNNNNNNNNNNNNNNNNN | NNNNNNNNNNNNNNNNNNNN | 1 |
| A | 4685 | hCoV-19/England/201360198/2020\|EPI_ISL_423236\|2020 | ···················· | ···················· | ···················· | ···················· | ···················· | NNNNNNNNNN·NNNN·N··· | 1 |
| A | 4686 | hCoV-19/England/201360200/2020\|EPI_ISL_423237\|2020 | ···················· | ···················· | ···················· | ···················· | ···················· | NNNNNNNNNNNNNNNNNNNN | 1 |
| A | 4688 | hCoV-19/England/201360202/2020\|EPI_ISL_423239\|2020 | ···················· | ···················· | ···················· | ···················· | ·NNNNNNNNNNNNNNNNNNN | NNNNNNNNNNNNNNN··NNN | 1 |
| A | 4695 | hCoV-19/England/201360217/2020\|EPI_ISL_423248\|2020 | ···················· | ···················· | ···················· | ···················· | NNNNNNNNNNNNNNNNNNNN | NNNNNNNNNNNNNNNNNNNN | 1 |
| A | 4697 | hCoV-19/England/20132080104/2020\|EPI_ISL_423804\|20 | ···················· | ···················· | ···················· | ···················· | ··NNNNNNNNNNNNNNNNNN | NNNNNNNNNNNNNNNNN··· | 1 |
| A | 4700 | hCoV-19/England/20132081304/2020\|EPI_ISL_423810\|20 | ···················· | ···················· | ···················· | ···················· | ··NNNNNNNNNNNNNNNNNN | ··NNN·N············· | 1 |
| A | 4701 | hCoV-19/England/20132081404/2020\|EPI_ISL_423811\|20 | ···················· | ···················· | ···················· | ···················· | ·NNNNNNNNNNNNNNNNNNN | NNNNNNNNNNNNNNNNNNNN | 1 |
| A | 4702 | hCoV-19/England/20132081804/2020\|EPI_ISL_423812\|20 | ···················· | ···················· | ···················· | ···················· | ··NNNNNNNNNNNNNNNNNN | NNNNNNNNNNNNNNNNNNNN | 1 |
| A | 4707 | hCoV-19/England/20132091504/2020\|EPI_ISL_423822\|20 | ···················· | ···················· | ···················· | ···················· | ·NNNNNNNNNNNNNNNNNNN | NNNNNNNNNNNNNNNNNNNN | 1 |
| A | 4714 | hCoV-19/England/20132099304/2020\|EPI_ISL_423832\|20 | ···················· | ···················· | ···················· | ···················· | NNNNNNNNNNNNNNNNNNNN | NNNNNNNNNNNNNNNNNNNN | 1 |
| A | 4724 | hCoV-19/England/20132108504/2020\|EPI_ISL_423851\|20 | ···················· | ···················· | ···················· | ···················· | NNNNNNNNNNNNNNNNNNNN | ·NN·N·N·N··N····NNNN | 1 |
| A | 4729 | hCoV-19/England/20126000604/2020\|EPI_ISL_423862\|20 | ···················· | ···················· | ···················· | ···················· | ·NNNNNNNNNNNNNNNNNNN | ··N················· | 1 |
| A | 4790 | hCoV-19/Scotland/EDB018/2020\|EPI_ISL_425824\|2020-0 | ···················· | ···················· | ···················· | ···················· | NNNNNNNNNNNNNNNNNNNN | NN·N···N·····NN···N· | 1 |
| A | 4797 | hCoV-19/Scotland/EDB041/2020\|EPI_ISL_425846\|2020-0 | ···················· | ···················· | ···················· | ···················· | NNNNNNNNNNNNNNNNNNNN | NNNNNNNNNNNNNNNNNNNN | 1 |
| A | 4799 | hCoV-19/Scotland/EDB044/2020\|EPI_ISL_425848\|2020-0 | ···················· | ···················· | ···················· | ···················· | ··············N····· | N······NN····NN····· | 1 |
| A | 4804 | hCoV-19/Scotland/EDB051/2020\|EPI_ISL_425855\|2020-0 | ···················· | ···················· | ···················· | ···················· | NNNNNNNNNNNNNNNNNNNN | NNNNNNNNNNNNNNNNNNNN | 1 |
| A | 4806 | hCoV-19/Scotland/EDB056/2020\|EPI_ISL_425860\|2020-0 | ···················· | ···················· | ···················· | ···················· | NNNNNNNNNNNNNNNNNNNN | NNNNNNNNNNNNNNNNNNNN | 1 |
| A | 4811 | hCoV-19/Scotland/EDB066/2020\|EPI_ISL_425865\|2020-0 | ···················· | ···················· | ···················· | ···················· | ········N·····NN···· | NNNN···NN··N········ | 1 |
| A | 4817 | hCoV-19/Scotland/EDB074/2020\|EPI_ISL_425871\|2020-0 | ···················· | ···················· | ···················· | ···················· | NNNNNNNNNNNNNNNNNNNN | NNNNNNNNNNNNNNNNNNNN | 1 |
| A | 4820 | hCoV-19/Scotland/EDB082/2020\|EPI_ISL_425874\|2020-0 | ···················· | ···················· | ···················· | ···················· | NNNNNNNNNNNNNNNNNNNN | NNNNNNNNNNNNNNNNNNNN | 1 |
| A | 4829 | hCoV-19/Scotland/EDB092/2020\|EPI_ISL_425883\|2020-0 | ···················· | ···················· | ···················· | ···················· | NNNNNNNNNNNNNNNNNNNN | NNNNNNNNNNNNNNNNNNNN | 1 |
| A | 4830 | hCoV-19/Scotland/EDB093/2020\|EPI_ISL_425884\|2020-0 | ···················· | ···················· | ···················· | ···················· | NNNNNNNNNNNNNNNNNNNN | NNNNNNNNNNNNNNNNNNNN | 1 |
| A | 4832 | hCoV-19/Scotland/EDB096/2020\|EPI_ISL_425886\|2020-0 | ···················· | ···················A | ···················· | ···················· | NNNNNNNNNNNNNNNNNNNN | NNNNNNNNNNNNNNNNNNNN | 1 |
| A | 4836 | hCoV-19/Scotland/EDB100/2020\|EPI_ISL_425890\|2020-0 | ···················· | ···················· | ···················· | ···················· | NNNNNNNNNNNNNNNNNNNN | NNNNNNNNNNNNNNNNNNNN | 1 |
| A | 4840 | hCoV-19/Scotland/EDB106/2020\|EPI_ISL_425894\|2020-0 | ···················· | ···················· | ···················· | ···················· | ···················· | NN·····NN··········· | 1 |
| A | 4841 | hCoV-19/Scotland/EDB107/2020\|EPI_ISL_425895\|2020-0 | ···················· | ···················· | ···················· | ···················· | NNNNNNNNNNNNNNNNNNNN | NNNNNNNNNNNNNNNNNNNN | 1 |
| A | 4844 | hCoV-19/Scotland/EDB110/2020\|EPI_ISL_425898\|2020-0 | ···················· | ···················· | ···················· | ···················· | NNNNNNNNNNNNNNNNNNNN | NNNNNNNNNNNNNNNNNNNN | 1 |
| A | 4968 | hCoV-19/England/20126001704/2020\|EPI_ISL_423866\|20 | ···················· | ·A·················· | ···················· | ···················· | ···················· | NNNNNNNNNNNNNNN····· | 1 |
| A | 5116 | hCoV-19/Jordan/SR-035/2020\|EPI_ISL_429995\|2020-03- | ···················· | ···················· | ···················· | ···················· | ·NNNNNN············· | ···················· | 1 |
| A | 5207 | hCoV-19/USA/UT-011/2020\|EPI_ISL_417959\|2020-03-11 | ···················· | ···················· | ···················· | ···················· | NNNNNNNNNNNNNNNNNNNN | NNNNNNNNNNNNNNNNNNNN | 1 |
| A | 5298 | hCoV-19/England/20132032704/2020\|EPI_ISL_423773\|20 | ···················· | ···················· | ···················· | ···················· | ··NN·········NNN··N· | NNNN················ | 1 |
| A | 5301 | hCoV-19/England/20132035704/2020\|EPI_ISL_423778\|20 | ···················· | ···················· | ···················· | ···················· | NNNNNNNNNNNNNNNNNNNN | NNNNNNNNNNNNNNNNNNNN | 1 |
| A | 5305 | hCoV-19/England/20132047404/2020\|EPI_ISL_423783\|20 | ···················· | ···················· | ···················· | ···················· | ·NNNNNNNNNNNNNNNNNNN | NNNNNNNNNNNNNNNNNNNN | 1 |
| A | 5309 | hCoV-19/England/20132066804/2020\|EPI_ISL_423791\|20 | ···················· | ···················· | ···················· | ···················· | ·NNNNNNNNNNNNNNNNNNN | NNNNNNNNNNNNNNNNNNNN | 1 |
| A | 5318 | hCoV-19/England/20128009404/2020\|EPI_ISL_423928\|20 | ···················· | ···················· | ···················· | ···················· | ···················· | NNNNNNNNNNNNNNNNN··· | 1 |
| A | 5321 | hCoV-19/England/20128014404/2020\|EPI_ISL_423938\|20 | ···················· | ···················· | ···················· | ···················· | ···················· | NNNNNNNNNNNNNNNNNNNN | 1 |
| A | 5322 | hCoV-19/England/20128014806/2020\|EPI_ISL_423940\|20 | ···················· | ···················· | ···················· | ···················· | NNNNNNNNNNNNNNNNNNNN | NNNNNNNNNNNNNNNNNNNN | 1 |
| A | 5323 | hCoV-19/England/20128036304/2020\|EPI_ISL_423953\|20 | ···················· | ···················· | ···················· | ···················· | ··N·NNNN··NN········ | NNNNNNNNNNNNNNNNNNNN | 1 |
| A | 5324 | hCoV-19/England/20128037406/2020\|EPI_ISL_423956\|20 | ···················· | ···················· | ···················· | ···················· | ···················· | NNNNNNNNNNNNNNNNNNNN | 1 |
| A | 5516 | hCoV-19/England/20126102404/2020\|EPI_ISL_423910\|20 | ···················· | ···················· | ···················· | ···················· | ··NNNNNNNNNNNNNNNNNN | NN·················· | 1 |
| A | 5522 | hCoV-19/England/20128035406/2020\|EPI_ISL_423951\|20 | ···················· | ···················· | ···················· | ···················· | ·NNNNNNNNNNNNNNNNNNN | NNNNNNNNNNNNNNNNNNNN | 1 |
| A | 5526 | hCoV-19/England/20128094604/2020\|EPI_ISL_423967\|20 | ···················· | ···················· | ···················· | ···················· | NNNNNNNNNNNNNNNNNNNN | ···················· | 1 |
| A | 5528 | hCoV-19/England/20128095504/2020\|EPI_ISL_423970\|20 | ···················· | ···················· | ···················· | ···················· | NNNNNNNNNNNNNNNNNNNN | NNNNNNNNNNNNNNNNNNNN | 1 |
| A | 5680 | hCoV-19/England/SHEF-BFD09/2020\|EPI_ISL_416735\|202 | ···················· | ···················· | ···················· | ···················· | ···N·NNNN·NN··NN·NN· | NN·······N·········· | 1 |
| A | 5682 | hCoV-19/England/SHEF-BFD27/2020\|EPI_ISL_416737\|202 | ···················· | ···················· | ···················· | ···················· | NNNNNNNNNNNNNNNNNNNN | NNNNNNNNNNNNNNNNNNNN | 1 |
| A | 5964 | hCoV-19/Iceland/113/2020\|EPI_ISL_417781\|2020-03-11 | ···················· | ···················· | ···················· | ···················· | ········NN·········· | NNNNNNNNNNNNNNNNNNNN | 1 |
| A | 5990 | hCoV-19/England/20130049604/2020\|EPI_ISL_423619\|20 | ···················· | ···················· | ···················· | ···················· | NNNNNNNNNNNNNNNNNNNN | NNNNNNNNNNNNNNNNNNNN | 1 |
| A | 6061 | hCoV-19/Australia/VIC850/2020\|EPI_ISL_427110\|2020- | ···················· | ···················· | ···················· | ···················· | NNNNNNNNNNNNNNNNNNNN | ···················· | 1 |
| A | 6074 | hCoV-19/Australia/VIC870/2020\|EPI_ISL_427125\|2020- | ···················· | ···················· | ···················· | ···················· | NNNNNNN············· | NNNNNNNNNNNNNNNNNNNN | 1 |
| A | 6149 | hCoV-19/England/20100022706/2020\|EPI_ISL_415136\|20 | ···················· | ···················· | ···················· | ···················· | NNNNNNNNNNNNNNNNNNNN | NNNNNNNNNNNNNNNNNNNN | 1 |
| A | 6152 | hCoV-19/England/20102000906/2020\|EPI_ISL_415148\|20 | ···················· | ···················· | ···················· | ···················· | ···N··NN·N·········· | ···················· | 1 |
| A | 6153 | hCoV-19/England/20110003506/2020\|EPI_ISL_415150\|20 | ···················· | ···················· | ···················· | ···················· | ·········N··N··N··NN | NNNNNNNNNNNNNNNNNNNN | 1 |
| A | 6220 | hCoV-19/Iceland/74/2020\|EPI_ISL_417701\|2020-03-09 | ···················· | ···················· | ···················· | ···················· | ···················· | NNNNNNNNNNNNNNNNNNNN | 1 |
| A | 6221 | hCoV-19/Iceland/33/2020\|EPI_ISL_417702\|2020-03-03 | ···················· | ···················· | ···················· | ···················· | ···················· | NNNNNNNNNNNNNNNNNNNN | 1 |
| A | 6240 | hCoV-19/Iceland/75/2020\|EPI_ISL_417726\|2020-03-09 | ···················· | ···················· | ···················· | ···················· | ·······NNNNNNNNNNNNN | NNNNNNNNNNNNNNNNNNNN | 1 |
| A | 6260 | hCoV-19/Iceland/24/2020\|EPI_ISL_417747\|2020-03-11 | ···················· | ···················· | ···················· | ···················· | ·······NNNNNNNNNNNNN | NNNNNNNNNNNNNNNNNNNN | 1 |
| A | 6277 | hCoV-19/Iceland/50/2020\|EPI_ISL_417769\|2020-03-05 | ···················· | ···················· | ···················· | ···················· | ···················· | NNNNNNNNNNNNNNNNNNNN | 1 |
| A | 6302 | hCoV-19/Australia/VIC885/2020\|EPI_ISL_427138\|2020- | ···················· | ···················· | ···················· | ···················· | NNNNNNNNNNNNNNNNNNNN | ···················· | 1 |
| A | 6305 | hCoV-19/Australia/VIC892/2020\|EPI_ISL_427144\|2020- | ···················· | ···················· | ···················· | ···················· | NNNNNNNNNNNNNNNNNNNN | NNNNNNNNNNNNNNNNNNNN | 1 |
| A | 6307 | hCoV-19/Australia/VIC894/2020\|EPI_ISL_427146\|2020- | ···················· | ···················· | ···················· | ···················· | NNNNNNNNNNNNNNNNNNNN | ···················· | 1 |
| A | 6311 | hCoV-19/Australia/VIC916/2020\|EPI_ISL_427151\|2020- | ···················· | ···················· | ···················· | ···················· | ···················· | NNNNNNNNNNNNNNNNNNNN | 1 |
| A | 6312 | hCoV-19/Australia/VIC920/2020\|EPI_ISL_427153\|2020- | ···················· | ···················· | ···················· | ···················· | NNNNNNNNNNNNNNNNNNNN | NNNNNNNNNNNNNNNNNNNN | 1 |
| A | 6313 | hCoV-19/Australia/VIC927/2020\|EPI_ISL_427155\|2020- | ···················· | ···················· | ···················· | ···················· | NNNNNNNNNNNNNNNNNNNN | ···················· | 1 |
| A | 6314 | hCoV-19/Australia/VIC929/2020\|EPI_ISL_427157\|2020- | ···················· | ···················· | ···················· | ···················· | NNNNNNNNNNNNNNNNNNNN | NNNNNNNNNNNNNNNNNNNN | 1 |
| A | 6321 | hCoV-19/USA/CT-UW-4341/2020\|EPI_ISL_427165\|2020-03 | ···················· | ···················A | ···················· | ···················· | N·············NNN··· | ···················· | 1 |
| A | 6326 | hCoV-19/Guangdong/20SF316/2020\|EPI_ISL_428461\|2020 | ···················· | ·A·················· | ···················· | ···················· | ···················· | ·······NNNNNNNNNNNNN | 1 |
| A | 6345 | hCoV-19/India/KA-nimh-0116/2020\|EPI_ISL_428480\|202 | ···················· | NNNNNNNNNNNNNNNNNNNN | ···················· | ···················· | ···················· | ···················· | 1 |
| A | 6356 | hCoV-19/Brazil/MG-CV49/2020\|EPI_ISL_429702\|2020-03 | ···················· | ···················· | ···················· | NNNNNNNNNNNNNNNNNNNN | ···················· | ···················· | 1 |
| A | 6470 | hCoV-19/England/20144023104/2020\|EPI_ISL_423524\|20 | ···················· | ···················· | ···················· | ···················· | ·NNNNNNNNNNNNNNNNNNN | NNNNNNNNNNNNNNNNNNNN | 1 |
| A | 6471 | hCoV-19/England/20144023904/2020\|EPI_ISL_423529\|20 | ···················· | ···················· | ···················· | ···················· | ···················· | NNNNNN··NNNN········ | 1 |
| A | 6474 | hCoV-19/England/20144060204/2020\|EPI_ISL_423548\|20 | ···················· | ···················· | ···················· | ···················· | NNNNNNNNNNNNNNNNNNNN | NNNNNNNNNNNNNNNNNNNN | 1 |
| A | 6478 | hCoV-19/England/20144067104/2020\|EPI_ISL_423556\|20 | ···················· | ···················· | ···················· | ···················· | NNNNNNNNNNNNNNNNNNNN | NNNN················ | 1 |
| A | 6482 | hCoV-19/England/20144074604/2020\|EPI_ISL_423564\|20 | ···················· | ···················· | ···················· | ···················· | ···················· | NNNN················ | 1 |

| | | | | | | | | | |
|---|---|---|---|---|---|---|---|---|---|
| A | 6486 | hCoV-19/England/20146007004/2020\|EPI_ISL_423575\|20 | ···················· | ···················· | ···················· | ···················· | ·NNNNNNNNNNNNNNNNNNN | NNNNNNNNNNNNNNNNNNNN | 1 |
| A | 6487 | hCoV-19/England/20146008504/2020\|EPI_ISL_423576\|20 | ···················· | ···················· | ···················· | ···················· | NNNNNNNNNNNNNNNNNNNN | NNNNNNNNNNNNNNNNNNNN | 1 |
| A | 6490 | hCoV-19/England/20148002504/2020\|EPI_ISL_423586\|20 | ···················· | ···················· | ···················· | ···················· | NNNNNNNNNNNNNNNNNNNN | ·········N······N·N· | 1 |
| A | 6491 | hCoV-19/England/20148004704/2020\|EPI_ISL_423587\|20 | ···················· | ···················· | ···················· | ···················· | ·NNNNNNNNNNNNNNNNNNN | NNNNNNNNNNNNNNNNNNNN | 1 |
| A | 6493 | hCoV-19/England/20148006704/2020\|EPI_ISL_423589\|20 | ···················· | ···················· | ···················· | ···················· | NNNNNNNNNNNNNNNNNNNN | NNNNNNNNNNNNNNNNNNNN | 1 |
| A | 6495 | hCoV-19/England/20148044604/2020\|EPI_ISL_423592\|20 | ···················· | ···················· | ···················· | ···················· | ·NNNNNNNNNNNNNNNNNNN | NNNNNNNNNNNNNNNNNNNN | 1 |
| A | 6499 | hCoV-19/England/20132004704/2020\|EPI_ISL_423716\|20 | ···T················ | ···················· | ···················· | ···················· | ··NNNNN············· | ···················· | 1 |
| A | 6506 | hCoV-19/England/20132018604/2020\|EPI_ISL_423732\|20 | ···················· | ···················· | ···················· | ···················· | ·NNNNNNNNNNNNNNNNNNN | NNNNNNNNNNNNNNNNNNNN | 1 |
| A | 6510 | hCoV-19/England/20132020804/2020\|EPI_ISL_423740\|20 | ···················· | ···················· | ···················· | ···················· | ···················· | NNNNNNNNNN·N········ | 1 |
| A | 6717 | hCoV-19/Iceland/21/2020\|EPI_ISL_417828\|2020-03-02 | ···················· | ···················· | ···················· | ···················· | ·········N··NN··N·N· | NNNNNNNNNNNNNNNNNNNN | 1 |
| A | 6732 | hCoV-19/Iceland/34/2020\|EPI_ISL_417843\|2020-03-03 | ···················· | ···················· | ···················· | ···················· | ······NNNNNNNNNNNNNN | NNNNNNNNNNNNNNNNNNNN | 1 |
| A | 6736 | hCoV-19/Iceland/51/2020\|EPI_ISL_417849\|2020-03-05 | ···················· | ···················· | ···················· | ···················· | ···················· | NNNNNNNNNNNNNNNNNNNN | 1 |
| A | 6757 | hCoV-19/Iceland/95/2020\|EPI_ISL_417874\|2020-03-10 | ···················· | ···················· | ···················· | ···················· | ···················· | NNNNNNNNNNNNNNNN···· | 1 |
| A | 6758 | hCoV-19/Iceland/96/2020\|EPI_ISL_417875\|2020-03-10 | ···················· | ···················· | ···················· | ···················· | ···················· | NNNNNNNNNNNNNNNNNNNN | 1 |
| A | 6844 | hCoV-19/England/20130064004/2020\|EPI_ISL_423649\|20 | ···················· | ···················· | ···················· | ···················· | ·NNNNNNNNNNNNNNNNNNN | NNNNNNNNNNNNNNNNNNNN | 1 |
| A | 6847 | hCoV-19/England/20130072404/2020\|EPI_ISL_423666\|20 | ···················· | ···················· | ···················· | ···················· | ·NNNNNNNNNNNNNNNNNNN | NNNNNNNNNNNNNNNNNNNN | 1 |
| A | 6848 | hCoV-19/England/201300860/2020\|EPI_ISL_423670\|2020 | ···················· | ···················· | ···················· | ···················· | ·NNNNNNNNNNNNNNNNNNN | NNNNNNNNNNNNNNNNNNNN | 1 |
| A | 6851 | hCoV-19/England/20131102804/2020\|EPI_ISL_423688\|20 | ···················· | ···················· | ···················· | ···················· | ···················· | NNNNNNNNNNNNNNNNNNNN | 1 |
| A | 7220 | hCoV-19/Spain/VC-FISABIO-1/2020\|EPI_ISL_414495\|202 | ···················· | ···················· | ···················· | ···················· | NNNNNNNNNNNNNNNNNNNN | NNNNNNNNNNNNNNNNNNNN | 1 |
| A | 7221 | hCoV-19/Spain/VC-FISABIO-2/2020\|EPI_ISL_414496\|202 | ···················· | ···················· | ···················· | ···················· | NNNNNNNNNNNNNNNNNNNN | NNNNNNNNNNNNNNNNNNNN | 1 |
| A | 7227 | hCoV-19/England/SHEF-C0118/2020\|EPI_ISL_420258\|202 | ···················· | ···················· | ···················· | ···················· | ···N······NN··NN···· | NNNN···NNN····NN···· | 1 |
| A | 7498 | hCoV-19/England/SHEF-C0109/2020\|EPI_ISL_420290\|202 | ···················· | ···················· | ···················· | ···················· | NNNNNNNNNNNNNNNNNNNN | NNNNNNNNNNNNNNNNNNNN | 1 |
| A | 7569 | hCoV-19/England/20130024003/2020\|EPI_ISL_420492\|20 | ···················· | ···················· | ···················· | ···················· | ····NNNNNNNNNNNNNNNN | NNNNNNNNNNNNNNNNNNNN | 1 |
| A | 7608 | hCoV-19/England/201360170/2020\|EPI_ISL_421769\|2020 | ···················· | ···················· | ···················· | ···················· | ···················· | NNNNNNNNNNNNNNNNNNNN | 1 |
| A | 7624 | hCoV-19/England/201361021/2020\|EPI_ISL_421789\|2020 | ···················· | ···················· | ···················· | ···················· | ···················· | NNNNNNNNNNNNNNN···N· | 1 |
| A | 7627 | hCoV-19/England/201361041/2020\|EPI_ISL_421792\|2020 | ···················· | ···················· | ···················· | ···················· | ·NNNNNNNNNNNNNNNNNNN | ·····N·············· | 1 |
| A | 7633 | hCoV-19/England/201380035/2020\|EPI_ISL_421799\|2020 | ···················· | ···················· | ···················· | ···················· | ··NNNNNNNNNNNNNNNNNN | NNNNNNNNNNNNNNNNNN·N | 1 |
| A | 7635 | hCoV-19/England/CAMB-73A82/2020\|EPI_ISL_425273\|202 | ···················· | ···················· | ···················· | ···················· | NNNNNNNNNNNNNNNNNNNN | NNNNNNNNNNNNNNNNNNNN | 1 |
| A | 7824 | hCoV-19/England/CAMB-737A9/2020\|EPI_ISL_425237\|202 | ···················· | ···················· | ···················· | ···················· | NNNNNNNNNNNNNNNNNNNN | NNNNNNNNNNNNNNNNNNNN | 1 |
| A | 8240 | hCoV-19/Canada/BC_65034/2020\|EPI_ISL_415585\|2020-0 | ···················· | ···················· | ···················· | ···················· | NNNNNNNNNNNNNNNNNNNN | NNNNNNNNNNNNNNNNNNNN | 1 |
| A | 8241 | hCoV-19/Canada/BC_64686/2020\|EPI_ISL_415586\|2020-0 | ···················· | ···················· | ···················· | ···················· | NNNNNNNNNNNNNNNNNNNN | NNNNNNNNNNNNNNNNNNNN | 1 |
| A | 8243 | hCoV-19/Canada/BC_83163/2020\|EPI_ISL_415589\|2020-0 | ···················· | ···················· | ···················· | ···················· | NNNNNNNNNNNNNNNNNNNN | NNNNNNNNNNNNNNNNNNNN | 1 |
| A | 8547 | hCoV-19/England/SHEF-C036D/2020\|EPI_ISL_420206\|202 | ···················· | ···················· | ···················· | ···················· | ··············N····· | NN·N···NN··········· | 1 |
| A | 8548 | hCoV-19/England/SHEF-C02E8/2020\|EPI_ISL_420207\|202 | ···················· | ···················· | ···················· | ···················· | NNNNNNNNNNNNNNNNNNNN | NNNNNNNNNNNNNNNNNNNN | 1 |
| A | 8555 | hCoV-19/England/SHEF-C0330/2020\|EPI_ISL_420214\|202 | ···················· | ···················· | ···················· | ···················· | NNNNNNNNNNNNNNNNNNNN | NNNNNNNNNNNNNNNNNNNN | 1 |
| A | 8559 | hCoV-19/England/SHEF-C02D9/2020\|EPI_ISL_420218\|202 | ···················· | ···················· | ···················· | ···················· | ···N··········NN···· | ···N···NN·NN········ | 1 |
| A | 8748 | hCoV-19/Belgium/ULG-2971/2020\|EPI_ISL_416997\|2020- | ···················· | ···················· | ···················· | ···················· | NNNNNNNNNNNNNNNNNNNN | NNNNNNNNNNNNNNNNNNNN | 1 |
| A | 9029 | hCoV-19/Norway/2090/2020\|EPI_ISL_420153\|2020-03-16 | ···················· | ···················· | ···················· | ···················· | NNNNNNNNNNNNNNNNNNNN | ···············NNNNN | 1 |
| A | 9030 | hCoV-19/England/SHEF-C093E/2020\|EPI_ISL_420154\|202 | ···················· | ···················· | ···················· | ···················· | NNNNNNNNNNNNNNNNNNNN | NNNNNNNNNNNNNNNNNNNN | 1 |
| A | 9052 | hCoV-19/England/SHEF-C092F/2020\|EPI_ISL_420176\|202 | ···················· | ···················· | ···················· | ···················· | NNNNNNNNNNNNNNNNNNN· | ···N················ | 1 |
| A | 9059 | hCoV-19/England/SHEF-C0303/2020\|EPI_ISL_420183\|202 | ···················· | ···················· | ···················· | ···················· | NNNNNNNNNNNNNNNNNNNN | NNNNNNNNNNNNNNNNNNNN | 1 |
| A | 9063 | hCoV-19/England/SHEF-C0242/2020\|EPI_ISL_420187\|202 | ···················· | ···················· | ···················· | ···················· | NNNNNNNNNNNNNNNNNNNN | NNNNNNNNNNNNNNNNNNNN | 1 |
| AA | 52 | hCoV-19/USA/NM-DOH-2020616673/2020\|EPI_ISL_751825\| | ···················· | ·A·················· | ···················· | ···················· | ···················· | NNNNNNNNNNNNNNNNNNNN | 1 |
| AA | 97 | hCoV-19/Singapore/1482/2020\|EPI_ISL_754097\|2020-12 | ···················· | ·A·················· | ···················· | ···················· | NNNN················ | ···················· | 1 |
| AA | 112 | hCoV-19/USA/ME-HETL-J0112/2020\|EPI_ISL_755315\|2020 | ···················· | ···················· | ···················· | ···················· | ···················· | NNNNNNNNNN·········· | 1 |
| AA | 461 | hCoV-19/USA/CA-LACPHL-AF00034/2020\|EPI_ISL_754117\| | ···················· | ···················· | ···················· | ···················· | ·NNNNNNNNNNNNNNNNNNN | NNNNNNNNNNNNNNNNNNNN | 1 |
| AA | 465 | hCoV-19/USA/ME-HETL-J0187/2020\|EPI_ISL_755365\|2020 | ···················· | ···················· | ···················· | ···················· | NNNNNNNNNNNNNNNNNNNN | NNNNNNNNNNNNNNN··NNN | 1 |
| AA | 468 | hCoV-19/USA/ME-HETL-J0193/2020\|EPI_ISL_755368\|2020 | ···················· | ···················· | ···················· | ···················· | NNNNNNNNNNNNNNNNN··· | ···················· | 1 |
| AA | 469 | hCoV-19/USA/ME-HETL-J0194/2020\|EPI_ISL_755369\|2020 | ···················· | ·A·················· | ···················· | ···················· | NNNNNNNNNNNNNNNNNNNN | NNNNNNNNNNNNNNNNNNNN | 1 |
| AA | 607 | hCoV-19/USA/CA-LACPHL-AF00042/2020\|EPI_ISL_754122\| | ···················· | ···················· | ···················· | ···················· | ··NNNNNNNNNNNNNNNNNN | NNNNNNNNNNNNNNNNNNNN | 1 |
| AA | 714 | hCoV-19/USA/ME-HETL-J0324/2020\|EPI_ISL_755453\|2020 | ···················· | ···················· | ···················· | ···················· | ·············NNNN··· | ···················· | 1 |
| AA | 720 | hCoV-19/USA/ME-HETL-J0338/2020\|EPI_ISL_755466\|2020 | ···················· | ···················· | ···················· | ···················· | ···················· | NNNNNNNNNNNNNNNNN··· | 1 |
| AA | 1183 | hCoV-19/Belarus/MA-RII-MH11359S/2020\|EPI_ISL_75423 | ···················· | ·A·················· | ···················· | ···················· | NNNNNNNN············ | ···················· | 1 |
| AA | 1208 | hCoV-19/USA/ME-HETL-J0426/2020\|EPI_ISL_755528\|2020 | ···················· | ···················· | ···················· | ···················· | NNNN················ | ···················· | 1 |
| AA | 1212 | hCoV-19/USA/ME-HETL-J0432/2020\|EPI_ISL_755532\|2020 | ···················· | ···················· | ···················· | ···················· | NNNNNNNNNNNNNNNNNNN· | ···················· | 1 |
| AA | 2672 | hCoV-19/USA/WA-UW-46999/2020\|EPI_ISL_756217\|2020-1 | ···················· | ·A·················· | ···················· | ···················· | ··········NNNNNNNNN· | ···················· | 1 |
| AA | 4819 | hCoV-19/USA/IL-UW-2449/2020\|EPI_ISL_759724\|2020-12 | ···················· | ···················· | ···················· | ···················· | NNNNNNNNNNNNNNNNNNNN | NNNNNNNNNNNNNNNNNNNN | 1 |
| AA | 4821 | hCoV-19/USA/WI-UW-2451/2020\|EPI_ISL_759726\|2020-12 | ···················· | ···················· | ···················· | ···················· | NNNNNNNNNNNNNNNNNNNN | NNNNNNNNNNNNNNNNNNNN | 1 |
| AA | 5102 | hCoV-19/USA/WI-UW-2460/2020\|EPI_ISL_759735\|2020-12 | ···················· | ···················· | ···················· | ···················· | NNNNNNNNNNNNNNNNNNNN | NNNNNNNNNNNNNNNNNNNN | 1 |
| AA | 5103 | hCoV-19/USA/WI-UW-2464/2020\|EPI_ISL_759739\|2020-12 | ···················· | ···················· | ···················· | ···················· | NNNNNNNNNNNNNNNNNNNN | NNNNNNNNNNNNNNNNNNNN | 1 |
| AA | 5104 | hCoV-19/USA/WI-UW-2465/2020\|EPI_ISL_759740\|2020-12 | ···················· | ···················· | ···················· | ···················· | NNNNNNNNNNNNNNNNNNNN | NNNNNNNNNNNNNNNNNNNN | 1 |
| AA | 5947 | hCoV-19/Germany/DE-BE-ChVir-D1972-8263/2020\|EPI_IS | ···················· | ···················· | ···················· | ···················· | NNNNNNNNNNNNNNNNNNNN | NNNNNNNNNNNNNNNNNNNN | 1 |
| AA | 5960 | hCoV-19/Germany/DE-BE-ChVir-W2202-4448/2020\|EPI_IS | ···················· | ·A···A·············· | ···················· | ···················· | NNNNNNNNNNNNNNNNNNNN | ···················· | 1 |
| AA | 6278 | hCoV-19/Germany/DE-BE-ChVir-W1297-7420/2020\|EPI_IS | ···················· | ···················· | ···················· | ···················· | ···················· | ··········NNNNNNNNN· | 1 |
| AA | 6339 | hCoV-19/England/205141172/2020\|EPI_ISL_756139\|2020 | ···················· | ···················· | ···················· | ···················· | NNNNNNNNNNNNNNNNNNNN | ···················· | 1 |
| AA | 6341 | hCoV-19/England/205141177/2020\|EPI_ISL_756141\|2020 | ···················· | ···················· | ···················· | ···················· | NNNNNNNNNNNNNNNNNNNN | NNNNNNNNNNNNNNNNNNNN | 1 |
| AA | 6344 | hCoV-19/England/205141190/2020\|EPI_ISL_756144\|2020 | ···················· | ···················· | ···················· | ···················· | NNNNNNNNNNNNNNNNNNNN | ···················· | 1 |
| AA | 7177 | hCoV-19/South Korea/KDCA0148/2020\|EPI_ISL_747383\|2 | ···················· | ···················· | ···················· | ···················· | NNNNN··············· | ···················· | 1 |
| AA | 7790 | hCoV-19/USA/WI-UW-2390/2020\|EPI_ISL_754626\|2020-10 | ···················· | ···················· | ···················· | ···················· | NNNNNNNNNNNNNNNNNNNN | NNNNNNNNNNNNNNNNNNNN | 1 |
| AA | 8072 | hCoV-19/USA/WI-UW-2401/2020\|EPI_ISL_754636\|2020-12 | ···················· | ···················· | ···················· | ···················· | NNNNNNNNNNNNNNNNNNNN | NNNNNNNNNNNNNNNNNNNN | 1 |
| AA | 8073 | hCoV-19/USA/WI-UW-2402/2020\|EPI_ISL_754637\|2020-12 | ···················· | ···················· | ···················· | ···················· | NNNNNNNNNNNNNNNNNNNN | NNNNNNNNNNNNNNNNNNNN | 1 |
| AA | 8077 | hCoV-19/USA/WI-UW-2438/2020\|EPI_ISL_754641\|2020-12 | ···················· | ···················· | ···················· | ···················· | NNNNNNNNNNNNNNNNNNNN | NNNNNNNNNNNNNNNNNNNN | 1 |
| AA | 8079 | hCoV-19/USA/WI-UW-2448/2020\|EPI_ISL_754643\|2020-12 | ···················· | ···················· | ···················· | ···················· | NNNNNNNNNNNNNNNNNNNN | NNNNNNNNNNNNNNNNNNNN | 1 |
| AA | 8083 | hCoV-19/USA/WI-UW-2447/2020\|EPI_ISL_754647\|2020-12 | ···················· | ···················· | ···················· | ···················· | NNNNNNNNNNNNNNNNNNNN | NNNNNNNNNNNNNNNNNNNN | 1 |
| AA | 8085 | hCoV-19/USA/IL-UW-2393/2020\|EPI_ISL_754649\|2020-12 | ···················· | ···················· | ···················· | ···················· | NNNNNNNNNNNNNNNNNNNN | NNNNNNNNNNNNNNNNNNNN | 1 |
| AA | 8089 | hCoV-19/USA/WI-UW-2444/2020\|EPI_ISL_754653\|2020-12 | ···················· | ···················· | ···················· | ···················· | NNNNNNNNNNNNNNNNNNNN | NNNNNNNNNNNNNNNNNNNN | 1 |
| AA | 8090 | hCoV-19/USA/WI-UW-2394/2020\|EPI_ISL_754654\|2020-12 | ···················· | ···················· | ···················· | ···················· | NNNNNNNNNNNNNNNNNNNN | NNNNNNNNNNNNNNNNNNNN | 1 |
| AA | 8093 | hCoV-19/USA/OH-UW-2399/2020\|EPI_ISL_754657\|2020-12 | ···················· | ···················· | ···················· | ···················· | NNNNNNNNNNNNNNNNNNNN | NNNNNNNNNNNNNNNNNNNN | 1 |
| AA | 8096 | hCoV-19/USA/WI-UW-2405/2020\|EPI_ISL_754660\|2020-12 | ···················· | ···················· | ···················· | ···················· | NNNNNNNNNNNNNNNNNNNN | NNNNNNNNNNNNNNNNNNNN | 1 |
| AA | 8103 | hCoV-19/USA/OH-UW-2397/2020\|EPI_ISL_754667\|2020-12 | ···················· | ···················· | ···················· | ···················· | NNNNNNNNNNNNNNNNNNNN | NNNNNNNNNNNNNNNNNNNN | 1 |

```
AA   8104 hCoV-19/USA/OH-UW-2398/2020|EPI_ISL_754668|2020-12   ···················· ···················· ···················· ···················· NNNNNNNNNNNNNNNNNNNN NNNNNNNNNNNNNNNNNNNN   1
AA   8105 hCoV-19/USA/WI-UW-2406/2020|EPI_ISL_754669|2020-12   ···················· ···················· ···················· ···················· NNNNNNNNNNNNNNNNNNNN NNNNNNNNNNNNNNNNNNNN   1
AB   182 hCoV-19/USA/NY-Wadsworth-292842-01/2020|EPI_ISL_76   ···················· ·A·············NNNNN ···················· ···················· ···················· ····················   1
AB   337 hCoV-19/USA/CA-LACPHL-AF00072/2020|EPI_ISL_766009|   ···················· ···················· ···················· ···················· ···················· NNNNNNNNNNNNNNNNNNNN   1
AB   555 hCoV-19/USA/LA-BIE-SUS2/2020|EPI_ISL_767405|2020-0   ···················· ···················· ···················· ···················· NNNNNNNNNNNNNNNNNNNN NNNNNNNNNNNNNNNNNNNN   1
AB   662 hCoV-19/Ireland/LH-NVRL-MPA78448/2020|EPI_ISL_7687   ···················· ···················· ···················· ···················· NNNNNNNNNNNNNNNNNNNN NNNNNNNNNNNNNNNNNNNN   1
AB   731 hCoV-19/Germany/HH-hpi-p1464/2020|EPI_ISL_775977|2   ···················· ···················· ···················· ···················· NNNNNNNNNNNNNNNNNNNN NNNNNNNNNNNNNNNNNNNN   1
AB   892 hCoV-19/England/205221138/2020|EPI_ISL_766245|2020   ···················· ···················· ···················· ···················· NNNNNNNNNNNNNNNNNNNN NNNNNNNNNNNNNNNNNNNN   1
AB   894 hCoV-19/England/205040390/2020|EPI_ISL_766247|2020   ···················· ···················· ···················· ···················· ···················· NNNNNNNN·····NN·····   1
AB   975 hCoV-19/Germany/HH-hpi-p2459/2020|EPI_ISL_775921|2   ···················· ···················· ···················· ···················· NNNNNNNNNNNNNNNNNNNN NNNNNNNNNNNNNNNNNNNN   1
AB   985 hCoV-19/Germany/HH-hpi-p2321/2020|EPI_ISL_775936|2   ···················· ···················· ···················· ···················· NNNNNNNNNNNNNNNNNNNN NNNNNNNNNNNNNNNNNNNN   1
AB   988 hCoV-19/Germany/HH-hpi-p2438/2020|EPI_ISL_775939|2   ···················· ···················· ···················· ···················· NNNNNNNNNNNNNNNNNNNN NNNNNNNNNNNNNNNNNNNN   1
AB   1141 hCoV-19/England/205100460/2020|EPI_ISL_766250|2020   ···················· ···················· ···················· ···················· ·NNNNNNNNNNNNNNNN··· ····················   1
AB   1158 hCoV-19/England/204690983/2020|EPI_ISL_766269|2020   ···················· ···················· ···················· ···················· NNNNNNNNNNNNNNNNNNNN NNNNNNNNNNNNNNNNNNNN   1
AB   1160 hCoV-19/England/204720099/2020|EPI_ISL_766271|2020   ···················· ···················· ···················· ···················· NNNNNNNNNNNNNNNNNNNN NNNNNNNNNNNNNNNNNNNN   1
AB   1162 hCoV-19/England/204742479/2020|EPI_ISL_766273|2020   ···················· ···················· ···················· ···················· ···················· N····NNN············   1
AB   1163 hCoV-19/England/204761672/2020|EPI_ISL_766274|2020   ···················· ···················· ···················· ···················· NNNNNNNNNNNNNNNNNNNN NNNNNNNNNNNNNNNNNNNN   1
AB   1164 hCoV-19/England/204761679/2020|EPI_ISL_766275|2020   ···················· ···················· ···················· ···················· NNNNNNNNNNNNNNNNNNNN NNNNNNNNNNNNNNNNNNNN   1
AB   1167 hCoV-19/England/204881987/2020|EPI_ISL_766278|2020   ···················· ···················· ···················· ···················· NNNNNNNNNNNNNNNNNNNN NNNNNNNNNNNNNNNNNNNN   1
AB   1313 hCoV-19/Wales/PHWC-4A6123/2020|EPI_ISL_765002|2020   ···················· ···················· ···················· ···················· ···················· NNNNNNNNNNNNNNNNNNNN   1
AB   1329 hCoV-19/Wales/PHWC-4A628A/2020|EPI_ISL_765018|2020   ···················· ···················· ···················· ···················· NNNNNNNNNNNNNNNNNNNN NNNNNNNNNNNNNNNNNNNN   1
AB   1340 hCoV-19/Wales/PHWC-4A6378/2020|EPI_ISL_765029|2020   ···················· ···················· ···················· ···················· ···················· NNNNNNNNNNNNNNNN····   1
AB   1630 hCoV-19/Wales/PHWC-4A6545/2020|EPI_ISL_765046|2020   ···················· ···················· ···················· ···················· NNNNNNNNNNNNNNNNNNNN NNNNNNNNNNNNNNNNNNNN   1
AB   1648 hCoV-19/Wales/PHWC-4A66D9/2020|EPI_ISL_765064|2020   ···················· ···················· ···················· ···················· NNNNNNN············· ····················   1
AB   1650 hCoV-19/Wales/PHWC-4A66F7/2020|EPI_ISL_765066|2020   ···················· ···················· ···················· ···················· NNNNNNNNNNNNNNNNNNNN NNNNNNNNN···········   1
AB   1655 hCoV-19/Wales/PHWC-4A677C/2020|EPI_ISL_765071|2020   ···················· ···················· ···················· ···················· NNNNNNNNNNNNNNNNNNNN NNNNNNNNNNNNNNNNNNNN   1
AB   1663 hCoV-19/Wales/PHWC-4A6879/2020|EPI_ISL_765079|2020   ···················· ···················· ···················· ···················· NNNNNNNNNNNN········ ····················   1
AB   1668 hCoV-19/Wales/PHWC-4A691C/2020|EPI_ISL_765084|2020   ···················· ···················· ···················· ···················· NNNNNNNNNNNNNNNNNNNN ····················   1
AB   1672 hCoV-19/Wales/PHWC-4A6976/2020|EPI_ISL_765088|2020   ···················· ···················· ···················· ···················· NNNNNNNNNNNNNNNNNNNN NNNNNNNNNNNNNNN·NNNN   1
AB   1676 hCoV-19/Wales/PHWC-4A69EF/2020|EPI_ISL_765092|2020   ···················· ···················· ···················· ···················· ···················· NNNNNNNNNNNNNNN·····   1
AB   1677 hCoV-19/Wales/PHWC-4A69FE/2020|EPI_ISL_765093|2020   ···················· ···················· ···················· ···················· NNNNNNNNNNNN········ ····················   1
AB   1840 hCoV-19/Wales/PHWC-4A3D40/2020|EPI_ISL_764624|2020   ···················· ···················· ···················· ···················· NNNNNNNNNNNNNNNNNNNN ····················   1
AB   1852 hCoV-19/Wales/PHWC-4A3E6B/2020|EPI_ISL_764636|2020   ···················· ···················· ···················· ···················· NNNN················ ····················   1
AB   1864 hCoV-19/Wales/PHWC-4A3F86/2020|EPI_ISL_764648|2020   ···················· ···················· ···················· ···················· NNNNNNNNNNNNNNNNNNNN ····················   1
AB   1865 hCoV-19/Wales/PHWC-4A3F95/2020|EPI_ISL_764649|2020   ···················· ···················· ···················· ···················· NNNNNNNNNNNNNNNNNNNN ····················   1
AB   1867 hCoV-19/Wales/PHWC-4A3FE0/2020|EPI_ISL_764651|2020   ···················· ···················· ···················· ···················· NNNNNNNNNNNNNNNNNNNN NNNNNNNNNNNNNNN·····   1
AB   1869 hCoV-19/Wales/PHWC-4A4037/2020|EPI_ISL_764653|2020   ···················· ···················· ···················· ···················· NNNN················ ····················   1
AB   1870 hCoV-19/Wales/PHWC-4A4046/2020|EPI_ISL_764654|2020   ···················· ···················· ···················· ···················· NNNNNNNNNNNNNNNNNNNN ············NNNNNNNN   1
AB   1871 hCoV-19/Wales/PHWC-4A4055/2020|EPI_ISL_764655|2020   ···················· ···················· ···················· ···················· NNNNNNNNNNNNNNNNNNNN NNNNNNNNNNNNNNNNNNNN   1
AB   1872 hCoV-19/Wales/PHWC-4A4213/2020|EPI_ISL_764667|2020   ···················· ···················· ···················· ···················· NNNNNNNNNNNNNNNNNNNN NNNNNNNNNNNNNNNNNNNN   1
AB   1874 hCoV-19/Wales/PHWC-4A4231/2020|EPI_ISL_764669|2020   ···················· ···················· ···················· ···················· NNNNNNNNNNNNNNNNNNNN NNNNNNNNN···········   1
AB   1875 hCoV-19/Wales/PHWC-4A4240/2020|EPI_ISL_764670|2020   ···················· ···················· ···················· ···················· NNNNNNNNNNNNNNNNNNNN NNNNNNNNNNNNNNNNNNNN   1
AB   1876 hCoV-19/Wales/PHWC-4A425F/2020|EPI_ISL_764671|2020   ···················· ···················· ···················· ···················· NNNNNNNNNNNNNNNNNNNN NNNNNNNNNNNNNNNNNNNN   1
AB   1878 hCoV-19/Wales/PHWC-4A428C/2020|EPI_ISL_764673|2020   ···················· ···················· ···················· ···················· NNNNNNNNNNNNNNNNNNNN NNNNNNNNNNNNNNNNNNNN   1
AB   1879 hCoV-19/Wales/PHWC-4A429B/2020|EPI_ISL_764674|2020   ···················· ···················· ···················· ···················· NNNNNNNNNNNNNNNNNNNN NNNNNNNNNNNNNNNNNNNN   1
AB   1880 hCoV-19/Wales/PHWC-4A42AA/2020|EPI_ISL_764675|2020   ···················· ···················· ···················· ···················· NNNNNNNNNNNNNNNNNNNN NNNNNNNNNNNNNNNNNNNN   1
AB   1881 hCoV-19/Wales/PHWC-4A42C8/2020|EPI_ISL_764676|2020   ···················· ···················· ···················· ···················· ··NNNNNNNNNNNNNNNNNN NNNNNNNNNNNNNNN·····   1
AB   1885 hCoV-19/Wales/PHWC-4A4310/2020|EPI_ISL_764680|2020   ···················· ···················· ···················· ···················· NNNNNNNNNNNNNNNNNNNN NNNNNNNNNNNNNNNNNNNN   1
AB   1886 hCoV-19/Wales/PHWC-4A433E/2020|EPI_ISL_764681|2020   ···················· ···················· ···················· ···················· NNNNNNNNNNNNNNNNNNNN NNNNNNNNNNNNNNNNNNNN   1
AB   1892 hCoV-19/Wales/PHWC-4A43C5/2020|EPI_ISL_764687|2020   ···················· ···················· ···················· ···················· ···················· NNNNNNNNNNNNNNN·····   1
AB   1894 hCoV-19/Wales/PHWC-4A43E3/2020|EPI_ISL_764689|2020   ···················· ···················· ···················· ···················· ···················· NNNNNNNNNNNNNNN·····   1
AB   1896 hCoV-19/Wales/PHWC-4A440E/2020|EPI_ISL_764691|2020   ···················· ···················· ···················· ···················· NNNNNNNNNNNNNNNNNNNN ····················   1
AB   1897 hCoV-19/Wales/PHWC-4A442C/2020|EPI_ISL_764692|2020   ···················· ···················· ···················· ···················· NNNNNNNNNNNNNNNNNNNN NNNNNNNNNNNNNNNNNNNN   1
AB   1898 hCoV-19/Wales/PHWC-4A444A/2020|EPI_ISL_764693|2020   ···················· ···················· ···················· ···················· NNNNNNNNNNNNNNNNNNNN ····················   1
AB   1900 hCoV-19/Wales/PHWC-4A4468/2020|EPI_ISL_764695|2020   ···················· ···················· ···················· ···················· ···················· NNNNNNNNNNNNNNN·····   1
AB   1901 hCoV-19/Wales/PHWC-4A4486/2020|EPI_ISL_764696|2020   ···················· ···················· ···················· ···················· NNNN················ ····················   1
AB   1902 hCoV-19/Wales/PHWC-4A4495/2020|EPI_ISL_764697|2020   ···················· ···················· ···················· ···················· NNNNNNNNNNNN········ NNNNNN··············   1
AB   1903 hCoV-19/Wales/PHWC-4A44A4/2020|EPI_ISL_764698|2020   ···················· ···················· ···················· ···················· NNNNNNNNNNNNNNNNNNNN ····················   1
AB   1918 hCoV-19/USA/WY-WPHL-20162034/2020|EPI_ISL_765973|2   ···················· ···················· ···················· ···················· NNNNNNNNNNNNNNNNNNNN ····················   1
AB   1999 hCoV-19/Israel/CVL-37-ngs/2020|EPI_ISL_776629|2020   ···················· ·A·················· ···················· ···················· ·····NNNNNNNNNNNNNNN NNNNNNNNNNNNNNNNNNN·   1
AB   2000 hCoV-19/Israel/CVL-48-ngs/2020|EPI_ISL_776635|2020   ···················· ·A·················· ···················· ···················· ·······NNNNNNNNNNNNN ····················   1
AB   2011 hCoV-19/Israel/CVL-73-ngs/2020|EPI_ISL_776646|2020   ···················· ···················· ···················· ···················· ··N····NNNNNNNNNNNNN ····················   1
AB   2013 hCoV-19/Israel/CVL-75-ngs/2020|EPI_ISL_776648|2020   ···················· ···················· ···················· ···················· NNNNNNNNNNNNNNNNN··· ····················   1
AB   2066 hCoV-19/Wales/PHWC-4A4152/2020|EPI_ISL_764658|2020   ···················· ···················· ···················· ···················· NNNNNNNNNNNNNNNNNNNN NNNNNNNNNNNNNNNNNNNN   1
AB   2067 hCoV-19/Wales/PHWC-4A4170/2020|EPI_ISL_764659|2020   ···················· ···················· ···················· ···················· NNNNNNNNNNNNNNNNNNNN NNNNNNNNNNNNNNNNNNNN   1
AB   2069 hCoV-19/Wales/PHWC-4A41AD/2020|EPI_ISL_764661|2020   ···················· ···················· ···················· ···················· NNNNNNNNNNNNNNNNNNNN NNNNNNNNNNNNNNNNNNNN   1
AB   2070 hCoV-19/Wales/PHWC-4A41BC/2020|EPI_ISL_764662|2020   ···················· ···················· ···················· ···················· ···················· NNNNNNNNNNNNNNNNNNNN   1
AB   2071 hCoV-19/Wales/PHWC-4A41CB/2020|EPI_ISL_764663|2020   ···················· ···················· ···················· ···················· ···················· NNNN·NNNNNNNNNN·····   1
AB   2072 hCoV-19/Wales/PHWC-4A41E9/2020|EPI_ISL_764664|2020   ···················· ···················· ···················· ···················· NNNNNNNNNNNNNNNNN··· ····················   1
AB   2073 hCoV-19/Wales/PHWC-4A41F8/2020|EPI_ISL_764665|2020   ···················· ···················· ···················· ···················· NNNN················ NNNNNNNNNNNNNNNNNNNN   1
AB   2075 hCoV-19/Wales/PHWC-4A44C2/2020|EPI_ISL_764700|2020   ···················· ···················· ···················· ···················· NNNNNNNNNNNNNNNNNNNN NNNNNNNNNNNNNNNNNNNN   1
AB   2076 hCoV-19/Wales/PHWC-4A44E0/2020|EPI_ISL_764701|2020   ···················· ·········A·········· ···················· ···················· NNNNNNNNNNNNNNNNNNNN NNNNNNNNNNNNNNNNNNNN   1
AB   2078 hCoV-19/Wales/PHWC-4A451A/2020|EPI_ISL_764703|2020   ···················· ···················· ···················· ···················· NNNNNNNNNNNNNNNNNNNN NNNNNNNNNNNNNNN·····   1
AB   2080 hCoV-19/Wales/PHWC-4A4538/2020|EPI_ISL_764705|2020   ···················· ···················· ···················· ···················· NNNNNNNNNNNNNNNNNNNN NNNNNNNNNNNNNNNNNNNN   1
AB   2081 hCoV-19/Wales/PHWC-4A4565/2020|EPI_ISL_764706|2020   ···················· ···················· ···················· ···················· NNNNNNNNNNNNNNNNNNNN NNNNNNNNNNNNNNNNNNNN   1
AB   2083 hCoV-19/Wales/PHWC-4A4583/2020|EPI_ISL_764708|2020   ···················· ···················· ···················· ···················· NNNNNNNNNNNNNNNNNNNN ····················   1
AB   2086 hCoV-19/Wales/PHWC-4A469F/2020|EPI_ISL_764720|2020   ···················· ···················· ···················· ···················· NNNNNNNNN··········· ····················   1
AB   2087 hCoV-19/Wales/PHWC-4A46AE/2020|EPI_ISL_764721|2020   ·················T·· ···················· ···················· ···················· NNNNNNNNNNNNNNNNNNNN NNNNNNNNNNNNNNNNNNNN   1
AB   2097 hCoV-19/Switzerland/SO-ETHZ-420319/2020|EPI_ISL_76   ···················· ···················· ···················· ···················· ·······NNNNNNNNNNNNN NNNNNNNNNNNNNNNNNNNN   1
AB   2102 hCoV-19/Switzerland/SO-ETHZ-420317/2020|EPI_ISL_76   ···················· ···················· ···················· ···················· ·······NNNNNNNNNNNNN NNNNNNNNNNNNNNNNNNNN   1
```

| | | | | | | | | |
|---|---|---|---|---|---|---|---|---|
| AB | 2103 hCoV-19/Switzerland/SZ-ETHZ-420324/2020\|EPI_ISL_76 | ···················· | ···················· | ···················· | ···················· | ·······NNNNNNNNNNNNN | ···················· | 1 |
| AB | 2104 hCoV-19/Switzerland/SZ-ETHZ-420323/2020\|EPI_ISL_76 | ···················· | ···················· | ···················· | ···················· | ·······NNNNNNNNNNNNN | NNNNNNNNNNNNNNNNNNNN | 1 |
| AB | 2105 hCoV-19/Switzerland/VS-ETHZ-420763/2020\|EPI_ISL_76 | ···················· | ···················· | ···················· | ···················· | ·······NNNNNNNNNNNNN | ···················· | 1 |
| AB | 2323 hCoV-19/Wales/PHWC-4A45DE/2020\|EPI_ISL_764712\|2020 | ···················· | ···················· | ···················· | ···················· | NNNNNNNNNNNNNNNNNNNN | ···················· | 1 |
| AB | 2325 hCoV-19/Wales/PHWC-4A4608/2020\|EPI_ISL_764714\|2020 | ···················· | ···················· | ···················· | ···················· | NNNNNNNNNNNNNNNNNNNN | NNNNNNNNNNNNNNNNNNNN | 1 |
| AB | 2326 hCoV-19/Wales/PHWC-4A4617/2020\|EPI_ISL_764715\|2020 | ···················· | ···················· | ···················· | ···················· | NNNNNNNNNNNNNNNNNNNN | NNNNNNNNNNNNNNNNNNNN | 1 |
| AB | 2329 hCoV-19/Wales/PHWC-4A4662/2020\|EPI_ISL_764718\|2020 | ···················· | ···················· | ···················· | ···················· | NNNNNNNNNNNNNNNNNNNN | NNNNNNNNNNNNNNNNNNNN | 1 |
| AB | 2330 hCoV-19/Wales/PHWC-4A4671/2020\|EPI_ISL_764719\|2020 | ···················· | ···················· | ···················· | ···················· | NNNNNNNNNNNNNNNNNNNN | ···················· | 1 |
| AB | 2331 hCoV-19/Wales/PHWC-4A46CC/2020\|EPI_ISL_764722\|2020 | ···················· | ···················· | ···················· | ···················· | ···················· | NNNN················ | 1 |
| AB | 2332 hCoV-19/Wales/PHWC-4A46EA/2020\|EPI_ISL_764723\|2020 | ···················· | ···················· | ···················· | ···················· | NNNNNNNNNNNNNNNNNNNN | NNNNNNNNNNNNNNNNNNNN | 1 |
| AB | 2336 hCoV-19/Wales/PHWC-4A4741/2020\|EPI_ISL_764727\|2020 | ···················· | ···················· | ···················· | ···················· | NNNNNNNNNNNNN······· | NNNN················ | 1 |
| AB | 2340 hCoV-19/Wales/PHWC-4A47C9/2020\|EPI_ISL_764731\|2020 | ···················· | ···················· | ···················· | ···················· | NNNNNNNNNNNNNNNNNNNN | ·······NNNNNNNNNNNNN | 1 |
| AB | 2345 hCoV-19/Wales/PHWC-4A4820/2020\|EPI_ISL_764736\|2020 | ···················· | ···················· | ···················· | ···················· | NNNNNNNNNNNNNNNNNNNN | NNNNNNNNNNNNNNNNNNNN | 1 |
| AB | 2347 hCoV-19/Wales/PHWC-4A484E/2020\|EPI_ISL_764738\|2020 | ···················· | ···················· | ···················· | ···················· | NNNNNNNNNNNNNNNNNNNN | ···················· | 1 |
| AB | 2355 hCoV-19/Wales/PHWC-4A490F/2020\|EPI_ISL_764746\|2020 | ···················· | ···················· | ···················· | ···················· | NNNNNNNNNNNNNNNNNNNN | NNNNNNNNNNNNNNNNNNNN | 1 |
| AB | 2356 hCoV-19/Wales/PHWC-4A491E/2020\|EPI_ISL_764747\|2020 | ···················· | ···················· | ···················· | ···················· | NNNNNNNNNNNNNNNNNNNN | NNNNNNNNNNNNNNNNNNNN | 1 |
| AB | 2363 hCoV-19/Wales/PHWC-4A49D2/2020\|EPI_ISL_764754\|2020 | ···················· | ···················· | ···················· | ···················· | NNNNNNNNNNNNNNNNNNNN | NNNNNNNNNNNNNNNNNNNN | 1 |
| AB | 2365 hCoV-19/Wales/PHWC-4A49F0/2020\|EPI_ISL_764756\|2020 | ···················· | ···················· | ···················· | ···················· | NNNNNNNNNNNNNNNNNNNN | NNNNNNNNNNNNNNNNNNNN | 1 |
| AB | 2378 hCoV-19/Wales/PHWC-4A4B18/2020\|EPI_ISL_764769\|2020 | ···················· | ···················· | ···················· | ···················· | NNNN·········NNN···· | NNNNNNNNNNNNNNN····· | 1 |
| AB | 2380 hCoV-19/Wales/PHWC-4A4B72/2020\|EPI_ISL_764771\|2020 | ···················· | ···················· | ···················· | ···················· | NNNNNNNNNNNNNNNNNNNN | NNNNNNNNNNNNNNNNNNNN | 1 |
| AB | 2381 hCoV-19/Wales/PHWC-4A4B90/2020\|EPI_ISL_764772\|2020 | ···················· | ···················· | ···················· | ···················· | NNNNNNNNNNNNNNNNNNNN | NNNNNNNNNNNNNNNNNNNN | 1 |
| AB | 2383 hCoV-19/Wales/PHWC-4A4BDC/2020\|EPI_ISL_764774\|2020 | ···················· | ···················· | ···················· | ···················· | NNNNNNNNNNNNNNNNNNNN | NNNNNNNNNNNNNNNNNNNN | 1 |
| AB | 2384 hCoV-19/Wales/PHWC-4A4BEB/2020\|EPI_ISL_764775\|2020 | ···················· | ···················· | ···················· | ···················· | NNNNNNNNNNNNNNNNNNNN | NNNNNNNNNNNNNNNNNNNN | 1 |
| AB | 2385 hCoV-19/Wales/PHWC-4A4BFA/2020\|EPI_ISL_764776\|2020 | ···················· | ···················· | ···················· | ···················· | NNNNNNNNNNNNNNNNNNNN | NNNNNNNNNNNNNNN····· | 1 |
| AB | 2387 hCoV-19/Wales/PHWC-4A4D9A/2020\|EPI_ISL_764789\|2020 | ···················· | ···················· | ···················· | ···················· | NNNNNNNNNNNNNNNNNNNN | NNNNNNNNNNNNNNNNNNNN | 1 |
| AB | 2389 hCoV-19/Wales/PHWC-4A4DB8/2020\|EPI_ISL_764791\|2020 | ···················· | ···················· | ···················· | ···················· | NNNNNNNNNNNNNNNNNNNN | NNNNNNNNNNNNNNNNNNNN | 1 |
| AB | 2390 hCoV-19/Wales/PHWC-4A4DC7/2020\|EPI_ISL_764792\|2020 | ···················· | ···················· | ···················· | ···················· | NNNNNNNNNNNNNNNNNNNN | NNNNNNNNNNNNNNNNNNNN | 1 |
| AB | 2391 hCoV-19/Wales/PHWC-4A4DE5/2020\|EPI_ISL_764793\|2020 | ···················· | ···················· | ···················· | ···················· | NNNNNNNNNNNNNNNNNNNN | ············NNN····· | 1 |
| AB | 2392 hCoV-19/Wales/PHWC-4A4E1F/2020\|EPI_ISL_764794\|2020 | ···················· | ···················· | ···················· | ···················· | NNNNNNNNNNNNNNNNNNNN | NNNNNNNNNNNNNNNNNNNN | 1 |
| AB | 2393 hCoV-19/Wales/PHWC-4A4E2E/2020\|EPI_ISL_764795\|2020 | ···················· | ···················· | ···················· | ···················· | ···················· | NNNNNNNNNNNNNNN····· | 1 |
| AB | 2394 hCoV-19/Wales/PHWC-4A4E6A/2020\|EPI_ISL_764796\|2020 | ···················· | ···················· | ···················· | ···················· | NNNNNNNNNNNNNNNNNNNN | NNNNNNNNNNNNNNNNNNNN | 1 |
| AB | 2395 hCoV-19/Wales/PHWC-4A4E79/2020\|EPI_ISL_764797\|2020 | ···················· | ···················· | ···················· | ···················· | NNNNNNNNNNNNNNNNNNNN | NNNNNNNNNNNNNNNNNNNN | 1 |
| AB | 2396 hCoV-19/Wales/PHWC-4A4EB5/2020\|EPI_ISL_764798\|2020 | ···················· | ···················· | ···················· | ···················· | NNNNNNNNNNNNNNNNNNNN | NNNNNNNNNNNNNNNNNNNN | 1 |
| AB | 2397 hCoV-19/Wales/PHWC-4A4EF1/2020\|EPI_ISL_764799\|2020 | ···················· | ···················· | ···················· | ···················· | NNNNNNNNNNNNNNNNNNNN | NNNNNNNNNNNNNNNNNNNN | 1 |
| AB | 2398 hCoV-19/Switzerland/ZH-ETHZ-420689/2020\|EPI_ISL_76 | ···················· | ···················· | ···················· | ···················· | ·······NNNNNNNNNNNNN | NNNNNNNNNNNNNNNNNNNN | 1 |
| AB | 2399 hCoV-19/Switzerland/ZH-ETHZ-420578/2020\|EPI_ISL_76 | ···················· | ···················· | ···················· | ···················· | ·······NNNNNNNNNNNNN | ···················· | 1 |
| AB | 2401 hCoV-19/Switzerland/ZH-ETHZ-420136/2020\|EPI_ISL_76 | ···················· | ···················· | ···················· | ·············S······ | ·······NNNNNNNNNNNNN | NNNNNNNNNNNNNNNNNNNN | 1 |
| AB | 2403 hCoV-19/Switzerland/ZH-ETHZ-420636/2020\|EPI_ISL_76 | ···················· | ···················· | ···················· | ···················· | ·······NNNNNNNNNNNNN | NNNNNNNNNNNNNNNNNNNN | 1 |
| AB | 2547 hCoV-19/Spain/M-20239437/2020\|EPI_ISL_776772\|2020- | ···················· | ···················· | ···················· | ···················· | NNNNNNNNNNNNNNNNNNNN | NNNNNNNNNNNNNNNNNNNN | 1 |
| AB | 2553 hCoV-19/Israel/CVL-190-ngs/2020\|EPI_ISL_776792\|202 | ···················· | ···················· | ···················· | ···················· | NNNNNNNNNNNNNNNNNNNN | NNNNNNNNNNNNNNNNNNNN | 1 |
| AB | 2571 hCoV-19/Wales/PHWC-4A4C24/2020\|EPI_ISL_764777\|2020 | ···················· | ···················· | ···················· | ···················· | NNNNNNNNNNNNNNNNNNNN | NNNNNNNNNNNNNNNNNNNN | 1 |
| AB | 2575 hCoV-19/Wales/PHWC-4A4CCA/2020\|EPI_ISL_764781\|2020 | ···················· | ···················· | ···················· | ···················· | NNNNNNNNNNNNNNNNNNNN | NNNNNNNNNNNNNNNNNNNN | 1 |
| AB | 2576 hCoV-19/Wales/PHWC-4A4CE8/2020\|EPI_ISL_764782\|2020 | ···················· | ···················· | ···················· | ···················· | NNNNNNNNNNNNNNNNNNNN | NNNNNNNNNNNNNNNNNNNN | 1 |
| AB | 2577 hCoV-19/Wales/PHWC-4A4D03/2020\|EPI_ISL_764783\|2020 | ···················· | ···················· | ···················· | ···················· | NNNNNNNNNNNNNNNNNNNN | NNNNNNNNNNNNNNNNNNNN | 1 |
| AB | 2578 hCoV-19/Wales/PHWC-4A4D21/2020\|EPI_ISL_764784\|2020 | ···················· | ···················· | ···················· | ···················· | NNNNNNNNNNNNNNNNNNNN | ···················· | 1 |
| AB | 2580 hCoV-19/Wales/PHWC-4A4D6D/2020\|EPI_ISL_764786\|2020 | ···················· | ···················· | ···················· | ···················· | NNNNNNNNNNNNNNNNNNNN | NNNNNNNNNNNNNNNNNNNN | 1 |
| AB | 2581 hCoV-19/Wales/PHWC-4A4D7C/2020\|EPI_ISL_764787\|2020 | ···················· | ···················· | ···················· | ···················· | NNNNNNNNNNNNNNNNNNNN | NNNNNNNNNNNNNNNNNNNN | 1 |
| AB | 2716 hCoV-19/Russia/KAM-RII-MH10619S/2020\|EPI_ISL_77067 | ···················· | ···················· | ···················· | ···················· | ···················· | NNNNNNNNNN·········· | 1 |
| AB | 2718 hCoV-19/Russia/KAM-RII-MH10624S/2020\|EPI_ISL_77067 | ···················· | ···················· | ···················· | ···················· | ···················· | NNNNNNNNNNNNNNNNNNNN | 1 |
| AB | 2818 hCoV-19/Wales/PHWC-4A4F3A/2020\|EPI_ISL_764800\|2020 | ···················· | ···················· | ···················· | ···················· | NNNNNNNNNNNNNNNNNNNN | NNNNNNNNNNNNNNNNNNNN | 1 |
| AB | 2819 hCoV-19/Wales/PHWC-4A4F49/2020\|EPI_ISL_764801\|2020 | ···················· | ···················· | ···················· | ···················· | NNNNNNNNNNNNNNNNNNNN | NNNNNNNNNNNNNNNNNNNN | 1 |
| AB | 2820 hCoV-19/Wales/PHWC-4A4F58/2020\|EPI_ISL_764802\|2020 | ···················· | ···················· | ···················· | ···················· | NNNNNNNNNNNNNNNNNNNN | NNNNNNNNNNNNNNNNNNNN | 1 |
| AB | 2821 hCoV-19/Wales/PHWC-4A4F76/2020\|EPI_ISL_764803\|2020 | ···················· | ···················· | ···················· | ···················· | NNNNNNNNNNNNNNNNNNNN | NNNNNNNNNNNNNNNNNNNN | 1 |
| AB | 2829 hCoV-19/Wales/PHWC-4A5018/2020\|EPI_ISL_764811\|2020 | ···················· | ···················· | ···················· | ···················· | NNNNNNNNNNNNNNNNN··· | ···················· | 1 |
| AB | 2835 hCoV-19/Wales/PHWC-4A5072/2020\|EPI_ISL_764817\|2020 | ···················· | ···················· | ···················· | ···················· | ··················NN | NNNNNNNNNNNNNNNNNNNN | 1 |
| AB | 2840 hCoV-19/Wales/PHWC-4A5124/2020\|EPI_ISL_764824\|2020 | ···················· | ···················· | ···················· | ···················· | ···················· | NNNNNNNNN·····N····· | 1 |
| AB | 2842 hCoV-19/Wales/PHWC-4A5142/2020\|EPI_ISL_764826\|2020 | ···················· | ···················· | ···················· | ···················· | NNNNNNNNNNNNNNNNNNNN | NNNNNNNNNNNNNNNNNNNN | 1 |
| AB | 2846 hCoV-19/Wales/PHWC-4A518E/2020\|EPI_ISL_764830\|2020 | ···················· | ···················· | ···················· | ···················· | NNNNN·····NNNNNNNNNN | ···················· | 1 |
| AB | 2849 hCoV-19/Wales/PHWC-4A51BB/2020\|EPI_ISL_764833\|2020 | ···················· | ···················· | ···················· | ···················· | NNNNNNNNNNNNNNNNNNNN | NNNNNNNNNNNNNNNNNNNN | 1 |
| AB | 2862 hCoV-19/Wales/PHWC-4A52A9/2020\|EPI_ISL_764846\|2020 | ···················· | ···················· | ···················· | ···················· | NNNNNNNNNNNNNNNNNNNN | NNNNNNNNNNNNNNNNNNNN | 1 |
| AB | 2863 hCoV-19/Wales/PHWC-4A52B8/2020\|EPI_ISL_764847\|2020 | ···················· | ···················· | ···················· | ···················· | ···················· | N···············NNNN | 1 |
| AB | 2868 hCoV-19/Wales/PHWC-4A533D/2020\|EPI_ISL_764852\|2020 | ···················· | ···················· | ···················· | ···················· | NNNNNNNNNNNNNNNNNNNN | NNNNNNNNNNNNNNNNNNNN | 1 |
| AB | 2872 hCoV-19/Wales/PHWC-4A5379/2020\|EPI_ISL_764856\|2020 | ···················· | ···················· | ···················· | ···················· | NNNNNNNNNNNNNNNNNNNN | ···················· | 1 |
| AB | 2873 hCoV-19/Wales/PHWC-4A5388/2020\|EPI_ISL_764857\|2020 | ···················· | ···················· | ···················· | ···················· | NNNNNNNNNNNNNNNNNNNN | NNNNNNNNNNNNNNN····· | 1 |
| AB | 2876 hCoV-19/Wales/PHWC-4A53B5/2020\|EPI_ISL_764860\|2020 | ···················· | ···················· | ···················· | ···················· | NNNNNNNNNNNN········ | ···················· | 1 |
| AB | 2880 hCoV-19/Wales/PHWC-4A540D/2020\|EPI_ISL_764864\|2020 | ···················· | ···················· | ···················· | ···················· | NNNNNNNNNNNNNNNNNNNN | NNNNNNNN·NNNNNNNNNNN | 1 |
| AB | 3091 hCoV-19/Wales/PHWC-4A5090/2020\|EPI_ISL_764818\|2020 | ···················· | ···················· | ···················· | ···················· | ···················· | ·NNNNNNNNNNNNNNN···· | 1 |
| AB | 3093 hCoV-19/Wales/PHWC-4A542B/2020\|EPI_ISL_764865\|2020 | ···················· | ···················· | ···················· | ···················· | NNNNNNNNNNNNNNNNNNNN | ·····NNNN··········· | 1 |
| AB | 3106 hCoV-19/Wales/PHWC-4A550A/2020\|EPI_ISL_764878\|2020 | ···················· | ···················· | ···················· | ···················· | NNNN················ | ···················· | 1 |
| AB | 3114 hCoV-19/Wales/PHWC-4A5582/2020\|EPI_ISL_764886\|2020 | ···················· | ···················· | ···················· | ···················· | NNNN················ | ···················· | 1 |
| AB | 3118 hCoV-19/Wales/PHWC-4A55CE/2020\|EPI_ISL_764890\|2020 | ···················· | ···················· | ···················· | ···················· | NNNNNNNNNNNNNNNNNNNN | NNNNNNNNNNNNNNNNNNNN | 1 |
| AB | 3301 hCoV-19/Spain/M-20224207/2020\|EPI_ISL_776871\|2020- | ···················· | ···················· | ···················· | ···················· | NNNNNNNNNNNNNNNNNNNN | NNNNNNNNNNNNNNNNNNNN | 1 |
| AB | 3304 hCoV-19/Spain/M-20191593/2020\|EPI_ISL_776875\|2020- | ···················· | ···················· | ···················· | ···················· | NNNNNNNNNNNNNNNNNNNN | NNNNNNNNNNNNNNNNNNNN | 1 |
| AB | 3329 hCoV-19/Wales/PHWC-4A5995/2020\|EPI_ISL_764942\|2020 | ···················· | ···················· | ···················· | ···················· | NNNNNNNNNNNNNNNNNNNN | NNNNNNNNNNNNNNNNNNNN | 1 |
| AB | 3331 hCoV-19/Wales/PHWC-4A59B3/2020\|EPI_ISL_764944\|2020 | ···················· | ···················· | ···················· | ···················· | NNNNNNNNNNNNNNNNNNNN | NNNNNNNNNNNNNNN····· | 1 |
| AB | 3332 hCoV-19/Wales/PHWC-4A59C2/2020\|EPI_ISL_764945\|2020 | ···················· | ···················· | ···················· | ···················· | NNNNNNNNNNNNNNNNNNNN | NNNNNNNNNNNNNNN····· | 1 |
| AB | 3335 hCoV-19/Wales/PHWC-4A5A1A/2020\|EPI_ISL_764948\|2020 | ···················· | ···················· | ···················· | ···················· | NNNNNNNNNNNNNNNNNNNN | ···················· | 1 |
| AB | 3338 hCoV-19/Wales/PHWC-4A5A83/2020\|EPI_ISL_764951\|2020 | ···················· | ···················· | ···················· | ···················· | ···················· | ·······NNNNNNNN····· | 1 |
| AB | 3339 hCoV-19/Wales/PHWC-4A5AA1/2020\|EPI_ISL_764952\|2020 | ···················· | ···················· | ···················· | ···················· | NNNNNNNNNNNNNNNNNNNN | NNNNNNNNNNNNNNNNNNNN | 1 |
| AB | 3672 hCoV-19/Wales/PHWC-4A57F5/2020\|EPI_ISL_764924\|2020 | ···················· | ···················· | ···················· | ···················· | NNNN················ | NNNNNNNNNNNNNNN····· | 1 |

| | | | | | | | | |
|---|---|---|---|---|---|---|---|---|
| AB | 3686 hCoV-19/Wales/PHWC-4A591D/2020\|EPI_ISL_764938\|2020 | ···················· | ···················· | ···················· | ···················· | NNNNNNNNNNNNNNNNNNNN | ··············NNNNNN | 1 |
| AB | 3687 hCoV-19/Wales/PHWC-4A5959/2020\|EPI_ISL_764939\|2020 | ···················· | ···················· | ···················· | ···················· | NNNNNNNNNNNNNNNNNNNN | ·······NNNNNNNNNNNNN | 1 |
| AB | 3688 hCoV-19/Wales/PHWC-4A5977/2020\|EPI_ISL_764940\|2020 | ···················· | ···················· | ···················· | ···················· | NNNNNNNNNNNNNNNNNNNN | ·················NNN | 1 |
| AB | 3689 hCoV-19/Wales/PHWC-4A5986/2020\|EPI_ISL_764941\|2020 | ···················· | ···················· | ···················· | ···················· | NNNNNNNNNNNNNNNNNNNN | NNNNNNNNNNNNNNNNNNNN | 1 |
| AB | 3690 hCoV-19/Wales/PHWC-4A5AFC/2020\|EPI_ISL_764953\|2020 | ···················· | ···················· | ···················· | ···················· | NNNNNNNNNNNNNNNNNNNN | NNNNNNNNNNNNNNNNNNNN | 1 |
| AB | 3691 hCoV-19/Wales/PHWC-4A5B26/2020\|EPI_ISL_764954\|2020 | ···················· | ···················· | ···················· | ···················· | NNNNNNNNNNNNNNNNNNNN | NNNNNNNNNNNNNNNNNNNN | 1 |
| AB | 3694 hCoV-19/Wales/PHWC-4A5B80/2020\|EPI_ISL_764957\|2020 | ···················· | ···················· | ···················· | ···················· | NNNNNNNNNNNNNNNNNNNN | NNNNNNNNNNNNNNNNNNNN | 1 |
| AB | 3696 hCoV-19/Wales/PHWC-4A5C05/2020\|EPI_ISL_764959\|2020 | ···················· | ···················· | ···················· | ···················· | NNNNNNNNNNNNNNNNNNNN | NNNNNNNNNNNNNNNNNNNN | 1 |
| AB | 3698 hCoV-19/Wales/PHWC-4A5C23/2020\|EPI_ISL_764961\|2020 | ···················· | ···················· | ···················· | ···················· | NNNNNNNNNNNNNNNNNNNN | NNNNNNNNNNNNNNNNNNNN | 1 |
| AB | 3699 hCoV-19/Wales/PHWC-4A5C32/2020\|EPI_ISL_764962\|2020 | ···················· | ·········A·········· | ···················· | ···················· | NNNNNNNNNNNNNNNNNNNN | ·······NNNNNNNN····· | 1 |
| AB | 3701 hCoV-19/Wales/PHWC-4A5C6F/2020\|EPI_ISL_764964\|2020 | ···················· | ···················· | ···················· | ···················· | NNNNNNNNNNNNNNNNNNNN | NNNNNNNNNNNNNNNNNNNN | 1 |
| AB | 3703 hCoV-19/Wales/PHWC-4A5CD8/2020\|EPI_ISL_764966\|2020 | ···················· | ···················· | ···················· | ···················· | NNNNNNNNNNNNNNNN···N | ···················· | 1 |
| AB | 3705 hCoV-19/Wales/PHWC-4A5D02/2020\|EPI_ISL_764968\|2020 | ···················· | ···················· | ···················· | ···················· | NNNNNNNNNNNNNNNNNNNN | NNNNNNNNNNNNNNNNNNNN | 1 |
| AB | 3706 hCoV-19/Wales/PHWC-4A5D11/2020\|EPI_ISL_764969\|2020 | ···················· | ···················· | ···················· | ···················· | NNNNNNNNNNNNNNNNNNNN | NNNNNNNNNNNNNNNNNNNN | 1 |
| AB | 3708 hCoV-19/Wales/PHWC-4A5D3F/2020\|EPI_ISL_764971\|2020 | ···················· | ···················· | ···················· | ···················· | NNNNNNNNNNNNNNNNNNNN | NNNNNNNNNNNNNNNNNNNN | 1 |
| AB | 3711 hCoV-19/Wales/PHWC-4A5DC6/2020\|EPI_ISL_764974\|2020 | ···················· | ···················· | ···················· | ···················· | NNNNNNNNNNNNNNNNNNNN | NNNNNNNNNNNNNNNNNNNN | 1 |
| AB | 3712 hCoV-19/Wales/PHWC-4A5DD5/2020\|EPI_ISL_764975\|2020 | ···················· | ···················· | ···················· | ···················· | NNNNNNNNNNNNNNNNNNNN | NNNNNNNNNN·········· | 1 |
| AB | 3714 hCoV-19/Wales/PHWC-4A5E3C/2020\|EPI_ISL_764977\|2020 | ···················· | ···················· | ···················· | ···················· | NNNNNNNNNNNNNNNNNNNN | NNNNNNNNNNNNNNNNNNNN | 1 |
| AB | 3716 hCoV-19/Wales/PHWC-4A5E87/2020\|EPI_ISL_764979\|2020 | ···················· | ···················· | ···················· | ···················· | NNNNNNNNNNNNNNNNNNNN | NNNNNNNNNNNNNNNNNNNN | 1 |
| AB | 3717 hCoV-19/Wales/PHWC-4A5E96/2020\|EPI_ISL_764980\|2020 | ···················· | ···················· | ···················· | ···················· | NNNNNNNNNNNNNNNNNNNN | NNNNNNNNNNNNNNNNNNNN | 1 |
| AB | 3718 hCoV-19/Wales/PHWC-4A5EF0/2020\|EPI_ISL_764981\|2020 | ···················· | ···················· | ···················· | ···················· | NNNNNNNNNNNNNNNNNNNN | NNNNNNNNNNNNNNNNNNNN | 1 |
| AB | 3721 hCoV-19/Wales/PHWC-4A5F57/2020\|EPI_ISL_764984\|2020 | ···················· | ···················· | ···················· | ···················· | NNNNNNNNNNNNNNNNNNNN | NNNNNNNNNNNNNNNNNNNN | 1 |
| AB | 3727 hCoV-19/Wales/PHWC-4A6008/2020\|EPI_ISL_764990\|2020 | ···················· | ···················· | ···················· | ···················· | NNNNNNNNNNNNNNNNNNNN | ···················· | 1 |
| AB | 3731 hCoV-19/Wales/PHWC-4A6062/2020\|EPI_ISL_764994\|2020 | ···················· | ···················· | ···················· | ···················· | NNNNNNNNNNNNNNNNNNNN | NNNNNNNNNNNNNNNNNNNN | 1 |
| AB | 3734 hCoV-19/Wales/PHWC-4A60BD/2020\|EPI_ISL_764997\|2020 | ···················· | ···················· | ···················· | ···················· | NNNNNNNNNNNNNNNNNNNN | NNNNNNNNNNNNNNNNNNNN | 1 |
| AB | 3735 hCoV-19/Wales/PHWC-4A60DB/2020\|EPI_ISL_764998\|2020 | ···················· | ···················· | ···················· | ···················· | NNNNNNNNNNNNNNNNNNNN | NNNNNNNNNNNNNNNNNNNN | 1 |
| AB | 3736 hCoV-19/Wales/PHWC-4A60EA/2020\|EPI_ISL_764999\|2020 | ···················· | ···················· | ···················· | ···················· | NNNNNNNNNNNNNNNNNNNN | ···················· | 1 |
| AB | 4027 hCoV-19/USA/MN-MDH-2229/2020\|EPI_ISL_770803\|2020-1 | ···················· | ···················· | ···················· | ···················· | ··NNNN·············· | ···················· | 1 |
| AB | 4119 hCoV-19/Wales/PHWC-4A3A1C/2020\|EPI_ISL_764273\|2020 | ···················· | ···················· | ···················· | ···················· | ···················· | NNNNNNNNNNNNNNNNNNNN | 1 |
| AB | 4120 hCoV-19/Wales/PHWC-4A42B9/2020\|EPI_ISL_764274\|2020 | ···················· | ···················· | ···················· | ···················· | NNNNNNNNNNNNNNNNNNNN | NNNNNNNNNNNNNNNNNNNN | 1 |
| AB | 4121 hCoV-19/Wales/PHWC-4A526D/2020\|EPI_ISL_764275\|2020 | ···················· | ···················· | ···················· | ···················· | NNNNNNNNNNNNNNNNNNNN | ···················· | 1 |
| AB | 4331 hCoV-19/Canada/ON-PHL-20-03725/2020\|EPI_ISL_778850 | ···················· | ·A·················· | ···················· | ···················· | NNNN················ | ···················· | 1 |
| AB | 4392 hCoV-19/Wales/PHWC-4A4732/2020\|EPI_ISL_764244\|2020 | ···················· | ···················· | ···················· | ···················· | NNNNNNNNNNNNNNNNNNNN | NNNNNNNNNNNNNNNNNNNN | 1 |
| AB | 4402 hCoV-19/Wales/PHWC-4A4D5E/2020\|EPI_ISL_764255\|2020 | ···················· | ···················· | ···················· | ···················· | NNNNNNNNNNNNNNNNNNNN | NNNNNNNNNNNNNNNNNNNN | 1 |
| AB | 4423 hCoV-19/Italy/LAZ-INMI-131/2020\|EPI_ISL_765569\|202 | ···················· | ···················· | ···················· | ···················· | NNNNNNNNNNNNNNNNNNNN | ···················· | 1 |
| AB | 4430 hCoV-19/USA/CO-Broad-CMU_00114/2020\|EPI_ISL_765578 | ···················· | ···················· | ···················· | ···················· | NNNNNNNNNNNNNNNNNNNN | NNNNNNNNN··········· | 1 |
| AB | 4535 hCoV-19/Germany/HH-hpi-p1913/2020\|EPI_ISL_776244\|2 | ···················· | ···················· | ···················· | ···················· | NNNNNNNNNNNNNNNNNNNN | NNNNNNNNNNNNNNNNNNNN | 1 |
| AB | 4622 hCoV-19/Germany/SN-RKI-NP-0070/2020\|EPI_ISL_763010 | ···················· | ···················· | ···················· | ···················· | NNNNNNNNNNNNNNNNNNNN | NNNNNNNNNNNNNNNNNNNN | 1 |
| AB | 4954 hCoV-19/Scotland/EDB11335/2020\|EPI_ISL_764368\|2020 | ···················· | ···················· | ···················· | ···················· | NNNNNNNNNNNNNNNNNNNN | NNNNNNNNNNNNNNNNNNNN | 1 |
| AB | 4955 hCoV-19/Northern Ireland/NIRE-234B5B/2020\|EPI_ISL_ | ···················· | ···················· | ···················· | ···················· | NNNNNNNNNNNNNNNNNNNN | ···················· | 1 |
| AB | 4958 hCoV-19/Northern Ireland/NIRE-234CC1/2020\|EPI_ISL_ | ···················· | ···················· | ···················· | ···················· | NNN··········NNNNNNN | ···················· | 1 |
| AB | 5040 hCoV-19/Canada/ON-PHL-20-03762/2020\|EPI_ISL_775072 | ···················· | ·A·················· | ···················· | ···················· | NNNNNNN············· | ···A················ | 1 |
| AB | 5164 hCoV-19/England/ALDP-CB4254/2020\|EPI_ISL_760839\|20 | ···················· | ···················· | ···················· | ···················· | NNNNNNNNNNNNNNNNNNNN | NNNNNNNNNNNNNNNNNNNN | 1 |
| AB | 5203 hCoV-19/Germany/NW-RKI-NP-0003/2020\|EPI_ISL_763029 | ···················· | NNNNNNNNNNNNNNNNNNNN | ···················· | ···················· | ···················· | ···················· | 1 |
| AB | 5207 hCoV-19/Germany/BW-RKI-NP-0071/2020\|EPI_ISL_763039 | ···················· | ···················· | ···················· | ···················· | NNNNNNNNNNNNNNNNNNNN | NNNNNNNNNNNNNNNNNNNN | 1 |
| AB | 5211 hCoV-19/Germany/BW-RKI-NP-0039/2020\|EPI_ISL_763044 | ···················· | ···················· | ···················· | ···················· | NNNNNNNNNNNNNNNNNNNN | NNNNNNNNNNNNNNNNNNNN | 1 |
| AB | 5216 hCoV-19/Germany/BW-RKI-NP-0015/2020\|EPI_ISL_763049 | ···················· | NNNNNNNNNNNNNNNNNNNN | ···················· | ···················· | ···················· | ···················· | 1 |
| AB | 5232 hCoV-19/Scotland/EDB11330/2020\|EPI_ISL_764367\|2020 | ···················· | ···················· | ···················· | ···················· | NNNNNNNNNNNNNNNNNNNN | NNNNNNNNNNNNNNNNNNNN | 1 |
| AB | 5234 hCoV-19/Scotland/EDB11337/2020\|EPI_ISL_764370\|2020 | ···················· | ···················· | ···················· | ···················· | NNNNNNNNNNNNNNNNNNNN | NNNNNNNNNNNNNNNNNNNN | 1 |
| AB | 5299 hCoV-19/Canada/ON-PHL-20-03812/2020\|EPI_ISL_775023 | ···················· | ·A·················· | ···················· | ···················· | NNNNNNNNNNNNNNNNNNNN | ···A················ | 1 |
| AB | 5321 hCoV-19/Canada/ON-PHL-20-03777/2020\|EPI_ISL_775086 | ···················· | ···················· | ···················· | ···················· | NNNNNNNNNNNNNNNNNNNN | ···················· | 1 |
| AB | 5410 hCoV-19/Northern Ireland/NIRE-234E52/2020\|EPI_ISL_ | ···················· | ···················· | ···················· | ···················· | NNNNNNNNNNNNNNNNN··· | ···················· | 1 |
| AB | 5415 hCoV-19/Northern Ireland/NIRE-235000/2020\|EPI_ISL_ | ···················· | ···················· | ···················· | ···················· | NNNNNNNNNNNNNNNNNNNN | ···················· | 1 |
| AB | 5416 hCoV-19/Northern Ireland/NIRE-23504C/2020\|EPI_ISL_ | ···················· | ···················· | ···················· | ···················· | NNNNNNNNNNNNNNNNNNNN | NNNNNNNNNNNNNNNNNNNN | 1 |
| AB | 5426 hCoV-19/Northern Ireland/NIRE-235316/2020\|EPI_ISL_ | ···················· | ···················· | ···················· | ···················· | NNNNNNNNNNNNNNNNNNNN | NNNNNNNNNNNNNNNN···· | 1 |
| AB | 5431 hCoV-19/Northern Ireland/NIRE-235413/2020\|EPI_ISL_ | ···················· | ···················· | ···················· | ···················· | NNNNNNNNNNNNNNNNNNNN | ···················· | 1 |
| AB | 5433 hCoV-19/Northern Ireland/NIRE-23547D/2020\|EPI_ISL_ | ···················· | ···················· | ···················· | ···················· | NNNNNNNNNNNNNNNNNNNN | NNNNNNNNNNNNNNNN···· | 1 |
| AB | 5438 hCoV-19/Northern Ireland/NIRE-235501/2020\|EPI_ISL_ | ···················· | ···················· | ···················· | ···················· | NNNN················ | ···················· | 1 |
| AB | 5527 hCoV-19/USA/MA-MGH-03300/2020\|EPI_ISL_765777\|2020- | ···················· | ···················· | ···················· | ···················· | ···················· | ··············NNNNNN | 1 |
| AB | 5566 hCoV-19/Canada/ON-PHL-20-03745/2020\|EPI_ISL_775105 | ···················· | ·A·················· | ···················· | ···················· | NNNNNNNNNNNNNNNNNNNN | ···A················ | 1 |
| AB | 5743 hCoV-19/Northern Ireland/NIRE-235149/2020\|EPI_ISL_ | ···················· | ···················· | ···················· | ···················· | NNNNNNNNNNNNNNNNNNNN | ···················· | 1 |
| AB | 5747 hCoV-19/Northern Ireland/NIRE-235219/2020\|EPI_ISL_ | ···················· | ···················· | ···················· | ···················· | NNNNNNN············· | ···················· | 1 |
| AB | 5750 hCoV-19/Northern Ireland/NIRE-235246/2020\|EPI_ISL_ | ···················· | ···················· | ···················· | ···················· | NNNN················ | ···················· | 1 |
| AB | 5752 hCoV-19/Northern Ireland/NIRE-235282/2020\|EPI_ISL_ | ···················· | ···················· | ···················· | ···················· | NNNNNNNNNNNNN······· | ···················· | 1 |
| AB | 5790 hCoV-19/Canada/ON-PHL-20-03611/2020\|EPI_ISL_775119 | ···················· | ·A·················· | ···················· | ···················· | NNNNNNNNNNNNNNNNNNNN | ···A················ | 1 |
| AB | 5810 hCoV-19/Canada/ON-PHL-20-03678/2020\|EPI_ISL_775162 | ···················· | ···················· | ···················· | ···················· | NNNNNNN············· | ···················· | 1 |
| AB | 5837 hCoV-19/Canada/ON-PHL-20-03707/2020\|EPI_ISL_775189 | ···················· | ···················· | ···················· | ···················· | NNNNNNNNNNNNNN······ | ···················· | 1 |
| AB | 5844 hCoV-19/Canada/ON-PHL-20-03714/2020\|EPI_ISL_775196 | ···················· | ···················· | ···················· | ···················· | NNNNNNNNNNNNNNNNNNNN | NNNNNNNNNNNNNNN····· | 1 |
| AB | 5851 hCoV-19/Germany/HH-hpi-p2354/2020\|EPI_ISL_776449\|2 | ···················· | ···················· | ···················· | ···················· | NNNNNNNNNNNNNNNNNNNN | NNNNNNNNNNNNNNNNNNNN | 1 |
| AB | 5863 hCoV-19/Germany/HH-hpi-p2383/2020\|EPI_ISL_776461\|2 | ···················· | ···················· | ···················· | ···················· | NNNNNNNNNNNNNNNNNNNN | NNNNNNNNNNNNNNNNNNNN | 1 |
| AB | 5867 hCoV-19/Germany/HH-hpi-p2387/2020\|EPI_ISL_776465\|2 | ···················· | ···················· | ···················· | ···················· | NNNNNNNNNNNNNNNNNNNN | NNNNNNNNNNNNNNNNNNNN | 1 |
| AB | 6039 hCoV-19/Switzerland/BE-ETHZ-420316/2020\|EPI_ISL_76 | ···················· | ·A·················· | ···················· | ···················· | ·······NNNNNNNNNNNNN | NNNNNNNNNNNNNNNNNNNN | 1 |
| AB | 6040 hCoV-19/Switzerland/BE-ETHZ-420680/2020\|EPI_ISL_76 | ···················· | ···················· | ···················· | ···················· | ·······NNNNNNNNNNNNN | NNNNNNNNNNNNNNNNNNNN | 1 |
| AB | 6042 hCoV-19/Switzerland/BL-ETHZ-420528/2020\|EPI_ISL_76 | ···················· | ···················· | ···················· | ···················· | ········NNNNNNNNNNNN | ···················· | 1 |
| AB | 6043 hCoV-19/Switzerland/BL-ETHZ-420201/2020\|EPI_ISL_76 | ···················· | ···················· | ···················· | ···················· | ·······NNNNNNNNNNNNN | ···················· | 1 |
| AB | 6046 hCoV-19/Switzerland/BL-ETHZ-420387/2020\|EPI_ISL_76 | ···················· | ···················· | ···················· | ···················· | ·······SNNNNNNNNNNNN | ···················· | 1 |
| AB | 6047 hCoV-19/Switzerland/BL-ETHZ-420228/2020\|EPI_ISL_76 | ···················· | ···················· | ···················· | ···················· | ·······NNNNNNNNNNNNN | NNNNNNNNNNNNNNNNNNNN | 1 |
| AB | 6131 hCoV-19/Germany/HH-hpi-p2414/2020\|EPI_ISL_776485\|2 | ···················· | ···················· | ···················· | ···················· | NNNNNNNNNNNNNNNNNNNN | NNNNNNNNNNNNNNNNNNNN | 1 |
| AB | 6137 hCoV-19/Germany/HH-hpi-p2479/2020\|EPI_ISL_776537\|2 | ···················· | ···················· | ···················· | ···················· | NNNNNNNNNNNNNNNNNNNN | NNNNNNNNNNNNNNNNNNNN | 1 |
| AB | 6248 hCoV-19/Wales/PHWC-4A385E/2020\|EPI_ISL_764582\|2020 | ···················· | ···················· | ···················· | ···················· | NNNNNNNNNNNNNNNNNNNN | NNNNNNNNNNNNNNNNNNNN | 1 |

| | | | | | | | | | |
|---|---|---|---|---|---|---|---|---|---|
| AB | 6249 | hCoV-19/Wales/PHWC-4A387C/2020\|EPI_ISL_764583\|2020 | ···················· | ···················· | ···················· | ···················· | NNNNNNNNNNNNNNNNNNNN | NNNNNNNNNNNNNNNNNNNN | 1 |
| AB | 6253 | hCoV-19/Wales/PHWC-4A38D6/2020\|EPI_ISL_764587\|2020 | ···················· | ···················· | ···················· | ···················· | NNNNNNNNNNNNNNNNNNNN | NNNNNNNNNNNNNNNNNNNN | 1 |
| AB | 6254 | hCoV-19/Wales/PHWC-4A38F4/2020\|EPI_ISL_764588\|2020 | ···················· | ···················· | ···················· | ···················· | NNNNNNNNNNNNNNNNNNN· | NNNNNNNNNNNNNNNNN··· | 1 |
| AB | 6321 | hCoV-19/Germany/HH-hpi-p2434/2020\|EPI_ISL_776501\|2 | ···················· | ···················· | ···················· | ···················· | NNNNNNNNNNNNNNNNNNNN | NNNNNNNNNNNNNNNNNNNN | 1 |
| AB | 6326 | hCoV-19/Germany/HH-hpi-p2440/2020\|EPI_ISL_776506\|2 | ···················· | ···················· | ···················· | ···················· | NNNNNNNNNNNNNNNNNNNN | NNNNNNNNNNNNNNNNNNNN | 1 |
| AB | 6336 | hCoV-19/Germany/HH-hpi-p2461/2020\|EPI_ISL_776520\|2 | ···················· | ···················· | ···················· | ···················· | NNNNNNNNNNNNNNNNNNNN | NNNNNNNNNNNNNNNNNNNN | 1 |
| AB | 6338 | hCoV-19/Germany/HH-hpi-p2463/2020\|EPI_ISL_776522\|2 | ···················· | ···················· | ···················· | ···················· | NNNNNNNNNNNNNNNNNNNN | NNNNNNNNNNNNNNNNNNNN | 1 |
| AB | 6355 | hCoV-19/Germany/HH-hpi-p2486/2020\|EPI_ISL_776543\|2 | ···················· | ·A·················· | ···················· | ···················· | NNNNNNNNNNNNNNNNNNNN | NNNNNNNNNNNNNNNNNNNN | 1 |
| AB | 6365 | hCoV-19/Germany/NW-hpi-p1761/2020\|EPI_ISL_776553\|2 | ···················· | ···················· | ···················· | ···················· | NNNNNNNNNNNNNNNNNNNN | NNNNNNNNNNNNNNNNNNNN | 1 |
| AB | 6450 | hCoV-19/Wales/PHWC-4A39B5/2020\|EPI_ISL_764594\|2020 | ···················· | ···················· | ···················· | ···················· | NNNNNNNNNNNNNNNNNNNN | NNNNNNNNNNNNNNNNNNNN | 1 |
| AB | 6451 | hCoV-19/Wales/PHWC-4A39E2/2020\|EPI_ISL_764595\|2020 | ···················· | ···················· | ···················· | ···················· | NNNNNNNNNNNNNNNNNNNN | NNNNNNNNNNNNNNNNNNNN | 1 |
| AB | 6452 | hCoV-19/Wales/PHWC-4A3A3A/2020\|EPI_ISL_764596\|2020 | ···················· | ···················· | ···················· | ···················· | NNNNNNNNNNNNNNNNNNNN | NNNNNNNNNNNNNNNNNNNN | 1 |
| AB | 6454 | hCoV-19/Wales/PHWC-4A3A58/2020\|EPI_ISL_764598\|2020 | ···················· | ···················· | ···················· | ···················· | NNNNNNNNNNNNNNNNNNNN | NNNNNNNNNNNNNNNNNNNN | 1 |
| AB | 6455 | hCoV-19/Wales/PHWC-4A3A76/2020\|EPI_ISL_764599\|2020 | ···················· | ···················· | ···················· | ···················· | NNNNNNNNNNNNNNNNNNNN | NNNNNNNNNNNNNNNNNNNN | 1 |
| AB | 6457 | hCoV-19/Wales/PHWC-4A3B64/2020\|EPI_ISL_764601\|2020 | ···················· | ···················· | ···················· | ···················· | NNNNNNNNNNNNNNNNNNNN | NNNNNNNNNNNNNNNNNNNN | 1 |
| AB | 6459 | hCoV-19/Wales/PHWC-4A3B91/2020\|EPI_ISL_764603\|2020 | ···················· | ···················· | ···················· | ···················· | NNNNNNNNNNNNNNNNNNNN | NNNNNNNNNNNNNNNNNNNN | 1 |
| AB | 6460 | hCoV-19/Wales/PHWC-4A3BBF/2020\|EPI_ISL_764604\|2020 | ···················· | ···················· | ···················· | ···················· | NNNNNNNNNNNNNNNNNNNN | NNNNNNNNNNNNNNNNNNNN | 1 |
| AB | 6496 | hCoV-19/USA/AZ-WL-CVG00126/2020\|EPI_ISL_765942\|202 | ···················· | ···················· | ···················· | ···················· | NNNNNNNNNNNNNNNNNNNN | NNNNNNNNNNNNNNNNNNNN | 1 |
| AB | 6517 | hCoV-19/Switzerland/SG-ETHZ-420081/2020\|EPI_ISL_76 | ···················· | ···················· | ···················· | ···················· | ······NNNNNNNNNNNNNN | NNNNNNNNNNNNNNNNNNNN | 1 |
| AB | 6519 | hCoV-19/Switzerland/SG-ETHZ-420700/2020\|EPI_ISL_76 | ···················· | ···················· | ···················· | ···················· | ······NNNNNNNNNNNN·· | ···················· | 1 |
| AB | 6520 | hCoV-19/Switzerland/SH-ETHZ-420087/2020\|EPI_ISL_76 | ···················· | ···················· | ···················· | ···················· | ······NNNNNNNNNNNNNN | NNNNNNNNNNNNNNNNNNNN | 1 |
| AB | 6521 | hCoV-19/Switzerland/SO-ETHZ-420716/2020\|EPI_ISL_76 | ···················· | ···················· | ···················· | ···················· | ······NNNNNNNNNNNNNN | NNNNNNNNNNNNNNNNNNNN | 1 |
| AB | 6523 | hCoV-19/Switzerland/SO-ETHZ-420719/2020\|EPI_ISL_76 | ··········T········· | ···················· | ···················· | ···················· | ······NNNNNNNNNNNNNN | ···················· | 1 |
| AB | 6527 | hCoV-19/Switzerland/SO-ETHZ-420485/2020\|EPI_ISL_76 | ···················· | ···················· | ···················· | ···················· | ······NNNNNNNNNNNNNN | NNNNNNNNNNNNNNNNNNNN | 1 |
| AB | 6580 | hCoV-19/England/205300064/2020\|EPI_ISL_770462\|2020 | ···················· | ···················· | ···················· | ···················· | ·NNNNNNNNNNNNNNNNNNN | NNNNNNNNNNNNNNNNNNNN | 1 |
| AB | 6582 | hCoV-19/England/205320747/2020\|EPI_ISL_770469\|2020 | ···················· | ···················· | ···················· | ···················· | ·NNNNNNNNNNNNNNNNNNN | NNNNNNNNNNNNNNNNNNNN | 1 |
| AB | 6643 | hCoV-19/Germany/HH-hpi-p2449/2020\|EPI_ISL_776511\|2 | ···················· | ···················· | ···················· | ···················· | NNNNNNNNNNNNNNNNNNNN | NNNNNNNNNNNNNNNNNNNN | 1 |
| AB | 6671 | hCoV-19/Wales/PHWC-4A6ADD/2020\|EPI_ISL_765100\|2020 | ···················· | ···················· | ···················· | ···················· | NNNNNNNNNNNNNNNNNNNN | ···················· | 1 |
| AB | 6674 | hCoV-19/Wales/PHWC-4A6B25/2020\|EPI_ISL_765103\|2020 | ···················· | ···················· | ···················· | ···················· | NNNNNNNNNNNNNNNNNNNN | ···················· | 1 |
| AB | 6677 | hCoV-19/Wales/PHWC-4A6BCB/2020\|EPI_ISL_765106\|2020 | ···················· | ···················· | ···················· | ···················· | ···················· | NNNN················ | 1 |
| AB | 6691 | hCoV-19/England/PORT-2D5679/2020\|EPI_ISL_765124\|20 | ···················· | ···················· | ···················· | ···················· | NNNNNNNNNNNNNNNNNNNN | NNNNNNNNNNNNNNNNNNNN | 1 |
| AB | 6836 | hCoV-19/Ireland/D-NVRL-MPD46296/2020\|EPI_ISL_76775 | ···················· | ···················· | ···················· | ···················· | NNNNNNNNNNNNNNNNNNNN | NNNNNNNNNNNNNNNNNNNN | 1 |
| AB | 6846 | hCoV-19/Ireland/D-NVRL-MPJ54572/2020\|EPI_ISL_76776 | ···················· | ···················· | ···················· | ···················· | NNNNNNNNNNNNNNNNNNNN | NNNNNNNNNNNNNNNNNNNN | 1 |
| AB | 6855 | hCoV-19/Ireland/D-NVRL-MPZ75781/2020\|EPI_ISL_76777 | ···················· | ···················· | ···················· | ···················· | NNNNNNNNNNNNNNNNNNNN | NNNNNNNNNNNNNNNNNNNN | 1 |
| AB | 6856 | hCoV-19/Ireland/D-NVRL-MPS48411/2020\|EPI_ISL_76777 | ···················· | ···················· | ···················· | ···················· | NNNNNNNNNNNNNNNNNNNN | NNNNNNNNNNNNNNNNNNNN | 1 |
| AB | 6903 | hCoV-19/Spain/M-20157372/2020\|EPI_ISL_778445\|2020- | ···················· | ···················· | ···················· | ···················· | NNNNNNNNNNNNNNNNNNNN | NNNNNNNNNNNNNNNNNNNN | 1 |
| AB | 6973 | hCoV-19/Wales/PHWC-4A6C7D/2020\|EPI_ISL_765111\|2020 | ···················· | ···················· | ···················· | ···················· | NNNNNNNNNNNNNNNNNNNN | NNNNNNNNNNNNNNNNNNNN | 1 |
| AB | 7367 | hCoV-19/Bangladesh/DNAS_DRCIM_isl_MT-04/2020\|EPI_I | ···················· | ···················· | ···················· | ········NNNNNN······ | ···················· | ···················· | 1 |
| AB | 7368 | hCoV-19/Bangladesh/DNAS_DRCIM_isl_MT-05/2020\|EPI_I | ···················· | ···················· | ···················· | NNNNNNNNNNNNNNN····· | ···················· | ···················· | 1 |
| AB | 7516 | hCoV-19/Portugal/PT1568/2020\|EPI_ISL_765220\|2020-0 | ···················· | ···················· | ···················· | ···················· | NNNN·N·············· | ···················· | 1 |
| AB | 7659 | hCoV-19/England/ALDP-C8FEC7/2020\|EPI_ISL_777292\|20 | ···················· | ···················· | ···················· | ···················· | NNNNNNNNNNNNNNNNNNNN | NNNNNNNNNNNNNNNNNNNN | 1 |
| AB | 7704 | hCoV-19/Wales/PHWC-4A690D/2020\|EPI_ISL_764063\|2020 | ···················· | ···················· | ···················· | ···················· | NNNNNNNNNNNNNNNNNNNN | NNN················· | 1 |
| AB | 7705 | hCoV-19/Wales/PHWC-4A4C42/2020\|EPI_ISL_764064\|2020 | ···················· | ···················· | ···················· | ···················· | NNNNNNNNNNNNNNNNNNNN | NNNNNNNNNNNNNNNNNNNN | 1 |
| AB | 7797 | hCoV-19/Switzerland/AG-ETHZ-420539/2020\|EPI_ISL_76 | ···················· | ···················· | ···················· | ···················· | ······NNNNNNNNNNNNNN | NNNNNNNNNNNNNNNNNNNN | 1 |
| AB | 7798 | hCoV-19/Switzerland/AG-ETHZ-420521/2020\|EPI_ISL_76 | ···················· | ·····A·············· | ···················· | ···················· | ······NNNNNNNNNNNNNN | ···················· | 1 |
| AB | 7799 | hCoV-19/Switzerland/AG-ETHZ-420519/2020\|EPI_ISL_76 | ···················· | ···················· | ···················· | ···················· | ······NNNNNNNNNNNNNN | NNNNNNNNNNNNNNNNNNNN | 1 |
| AB | 7801 | hCoV-19/Switzerland/AG-ETHZ-420678/2020\|EPI_ISL_76 | ···················· | ···················· | ···················· | ···················· | ······NNNNNNNNNNNNNN | ···················· | 1 |
| AB | 7831 | hCoV-19/Germany/HH-hpi-p1511/2020\|EPI_ISL_776016\|2 | ···················· | ···················· | ···················· | ···················· | NNNNNNNNNNNNNNNNNNNN | NNNNNNNNNNNNNNNNNNNN | 1 |
| AB | 7846 | hCoV-19/Germany/HH-hpi-p1537/2020\|EPI_ISL_776031\|2 | ···················· | ···················· | ···················· | ···················· | NNNNNNNNNNNNNNNNNNNN | NNNNNNNNNNNNNNNNNNNN | 1 |
| AB | 8074 | hCoV-19/USA/CO-CDPHE-2011020168/2020\|EPI_ISL_77123 | ···················· | ···················· | ···················· | ···················· | NNNNNNNNNNNNNNNNNNNN | ···················· | 1 |
| AB | 8084 | hCoV-19/USA/CO-CDPHE-2011103380/2020\|EPI_ISL_77124 | ···················· | ···················· | ···················· | ···················· | NNNNNNNNNNNNNNNNNNNN | ···················· | 1 |
| AB | 8123 | hCoV-19/Germany/HH-hpi-p1581/2020\|EPI_ISL_776056\|2 | ···················· | ···················· | ···················· | ···················· | NNNNNNNNNNNNNNNNNNNN | NNNNNNNNNNNNNNNNNNNN | 1 |
| AB | 8215 | hCoV-19/England/PORT-2D55D6/2020\|EPI_ISL_764119\|20 | ···················· | ·A·················· | ···················· | ···················· | NNNNNNNNNNNNNNNNNNNN | NNNNNNNNNNNNNNNNNNNN | 1 |
| AB | 8223 | hCoV-19/Northern Ireland/NIRE-2352EC/2020\|EPI_ISL_ | ···················· | ···················· | ···················· | ···················· | NNNN················ | ···················· | 1 |
| AB | 8294 | hCoV-19/USA/CO-CDPHE-2100010210/2020\|EPI_ISL_77129 | ·······T············ | ···················· | ···················· | ···················· | NNNNNNNNNNNNNNNNNNNN | ···················· | 1 |
| AB | 8351 | hCoV-19/Germany/HH-hpi-p1796/2020\|EPI_ISL_776165\|2 | ···················· | ···················· | ···················· | ···················· | NNNNNNNNNNNNNNNNNNNN | NNNNNNNNNNNNNNNNNNNN | 1 |
| AB | 8376 | hCoV-19/Spain/M-20120611/2020\|EPI_ISL_777490\|2020- | ···················· | ···················· | ···················· | ···················· | NNNNNNNNNNNNNNNNNNNN | NNNNNNNNNNNNNNNNNNNN | 1 |
| AB | 8612 | hCoV-19/USA/CO-CDPHE-2100016524/2020\|EPI_ISL_77132 | ···················· | ···················· | ···················· | ···················· | NNNNNNNNNNNNNNNNNNNN | ···········NNN······ | 1 |
| AB | 8623 | hCoV-19/USA/CO-CDPHE-2100037007/2020\|EPI_ISL_77134 | ·······T············ | ·A·················· | ···················· | ···················· | NNNNNNNNNNNNNNNNNNNN | NNNNNNNNNNNNNNNNNNNN | 1 |
| AB | 8624 | hCoV-19/USA/CO-CDPHE-2100037611/2020\|EPI_ISL_77134 | ···················· | ·A·················· | ···················· | ···················· | NNNNNNNNNNNNNNNNNNNN | NN·········NNNN····· | 1 |
| AB | 8625 | hCoV-19/USA/CO-CDPHE-2100037717/2020\|EPI_ISL_77134 | ···················· | ···················· | ···················· | ···················· | NNNNNNNNNNNNNNNNNNNN | NNNNNNNNNNNNNNN····· | 1 |
| AB | 8629 | hCoV-19/USA/CO-CDPHE-2100042725/2020\|EPI_ISL_77135 | ···················· | ···················· | ···················· | ···················· | NNNNNNNNNNNNNNNNNNNN | ···················· | 1 |
| AB | 8734 | hCoV-19/USA/CO-CDPHE-2100028667/2020\|EPI_ISL_77133 | ···················· | ···················· | ···················· | ···················· | NNNNNNNNNNNNNNNNN··· | ···················· | 1 |
| B | 39 | hCoV-19/USA/WA-S302/2020\|EPI_ISL_430156\|2020-03-04 | ···················· | ···················· | NNNNN··············· | ···················· | ···················· | ···················· | 1 |
| B | 85 | hCoV-19/Spain/VC-FISABIO-202/2020\|EPI_ISL_436297\|2 | ···················· | ···················· | ···················· | ···················· | NNNNNNNNNNNNNNNNNNNN | ···················· | 1 |
| B | 265 | hCoV-19/Spain/VC-FISABIO-105/2020\|EPI_ISL_436200\|2 | ···················· | ···················· | ···················· | ···················· | NNNNNNNNNNNNNNNNNNNN | NNNNNNNNNNNNNNN····· | 1 |
| B | 267 | hCoV-19/Spain/VC-FISABIO-107/2020\|EPI_ISL_436202\|2 | ···················· | ···················· | ···················· | ···················· | NNNNNNNNNNNNNNNNNNNN | NNNNNNNNNNNNNNNNNNNN | 1 |
| B | 270 | hCoV-19/Spain/VC-FISABIO-111/2020\|EPI_ISL_436206\|2 | ···················· | ···················· | ···················· | ···················· | NNNNNNNNNNNN········ | ···················· | 1 |
| B | 271 | hCoV-19/Spain/VC-FISABIO-112/2020\|EPI_ISL_436207\|2 | ···················· | ···················· | ···················· | ···················· | ·NNNNNN············· | ···················· | 1 |
| B | 272 | hCoV-19/Spain/VC-FISABIO-114/2020\|EPI_ISL_436209\|2 | ···················· | ···················· | ···················· | ···················· | NNNNNNNNNNNNNNNNNNNN | ···················· | 1 |
| B | 278 | hCoV-19/Spain/VC-FISABIO-122/2020\|EPI_ISL_436217\|2 | ···················· | ···················· | ···················· | ···················· | NNNNNNNNNNNNNNNNNNNN | NNNNNNNNNNNNNNNNNNNN | 1 |
| B | 285 | hCoV-19/Spain/VC-FISABIO-134/2020\|EPI_ISL_436229\|2 | ···················· | ···················· | ···················· | ···················· | ··NN···NNNN········· | ···················· | 1 |
| B | 288 | hCoV-19/Spain/VC-FISABIO-138/2020\|EPI_ISL_436233\|2 | ···················· | ···················· | ···················· | ···················· | NNNNNNNNNNNNNNNNNNNN | NNNNNNNNNNNNNNN·NNNN | 1 |
| B | 289 | hCoV-19/Spain/VC-FISABIO-139/2020\|EPI_ISL_436234\|2 | ···················· | ···················· | ···················· | ···················· | NNNNNNNNNNNNNNNNNNNN | ···················· | 1 |
| B | 290 | hCoV-19/Spain/VC-FISABIO-140/2020\|EPI_ISL_436235\|2 | ···················· | ···················· | ···················· | ···················· | NNNNNNNNNNNNNNNNNNNN | NNNNNNNNNNNNNNNNNNNN | 1 |
| B | 292 | hCoV-19/Spain/VC-FISABIO-142/2020\|EPI_ISL_436237\|2 | ···················· | ···················· | ···················· | ···················· | ·NNNNNNNNNNNNNNNNNN· | ···················· | 1 |
| B | 294 | hCoV-19/Spain/VC-FISABIO-144/2020\|EPI_ISL_436239\|2 | ···················· | ···················· | ···················· | ···················· | NNNNNNNNNNNNNNNNNNNN | NNNNNNN····NNNN····N | 1 |
| B | 298 | hCoV-19/Spain/VC-FISABIO-149/2020\|EPI_ISL_436244\|2 | ···················· | ···················· | ···················· | ···················· | NNNNNNNNNNNNNNNNNNNN | NNNNNNNNNNNNNNN····· | 1 |
| B | 310 | hCoV-19/Spain/VC-FISABIO-166/2020\|EPI_ISL_436261\|2 | ···················· | ···················· | ···················· | ···················· | ·NNNNNN···NN········ | NNNNNNN············· | 1 |
| B | 312 | hCoV-19/Spain/VC-FISABIO-168/2020\|EPI_ISL_436263\|2 | ···················· | ···················· | ···················· | ···················· | NNNNNNNNNNNNNNNNNNNN | ···················· | 1 |

| | | | | | | | | | |
|---|---|---|---|---|---|---|---|---|---|
| B | 313 | hCoV-19/Spain/VC-FISABIO-169/2020\|EPI_ISL_436264\|2 | ···················· | ···················· | ···················· | ···················· | NNNNNNNNNNNNNNNNNNNN | NNNNNNNNNNNNN······· | 1 |
| B | 315 | hCoV-19/Spain/VC-FISABIO-171/2020\|EPI_ISL_436266\|2 | ···················· | ···················· | ···················· | ···················· | NNNNNNNNNNNNNNNNNNNN | NNNNNNNNNNNNNNNNNNNN | 1 |
| B | 316 | hCoV-19/Spain/VC-FISABIO-172/2020\|EPI_ISL_436267\|2 | ···················· | ···················· | ···················· | ···················· | ··NNNNN············· | ···················· | 1 |
| B | 398 | hCoV-19/Scotland/CVR1692/2020\|EPI_ISL_438845\|2020- | ···················· | ···················· | ···················· | ···················· | ······NNNNNNNNNNNNNN | ···················· | 1 |
| B | 577 | hCoV-19/USA/WA-S242/2020\|EPI_ISL_430280\|2020-03-01 | ···················· | ···················· | ···················· | ···················· | NNNNNNNNNNNNNNNNNNNN | ···················· | 1 |
| B | 598 | hCoV-19/Scotland/EDB912/2020\|EPI_ISL_432876\|2020-0 | ···················· | ···················· | ···················· | ···················· | NNNNNNNNNNNNNNNNNNNN | NNNNNNNNNNNNNNNNNNNN | 1 |
| B | 600 | hCoV-19/Scotland/EDB918/2020\|EPI_ISL_432878\|2020-0 | ···················· | ···················· | ···················· | ···················· | NNNNNNNNNNNNNNNNNNNN | NNNNNNNNNNNNNNNNNNNN | 1 |
| B | 606 | hCoV-19/Scotland/EDB929/2020\|EPI_ISL_432888\|2020-0 | ···················· | ···················· | ···················· | ···················· | NNNNNNNNNNNNNNNNNNNN | NNNNNNNNNNNNNNNNNNNN | 1 |
| B | 607 | hCoV-19/Scotland/EDB932/2020\|EPI_ISL_432889\|2020-0 | ···················· | ···················· | ···················· | ···················· | NNNNNNNNNNNNNNNNNNNN | NNNNNNNNNNNNNNNNNNNN | 1 |
| B | 614 | hCoV-19/Scotland/EDB996/2020\|EPI_ISL_432897\|2020-0 | ···················· | ·A·················· | ···················· | ···················· | NNNNNNNNNNNNNNNNNNNN | NNNNNNNNNNNNNNNNNNNN | 1 |
| B | 863 | hCoV-19/Spain/VC-FISABIO-230/2020\|EPI_ISL_436325\|2 | ···················· | ···················· | ···················· | ···················· | NNNNNNNNNNNNNNNNNNNN | ···················· | 1 |
| B | 869 | hCoV-19/Spain/VC-FISABIO-241/2020\|EPI_ISL_436336\|2 | ···················· | ···················· | ···················· | ···················· | NNNNNNNNNNNNNNNNNNNN | ···················· | 1 |
| B | 871 | hCoV-19/Spain/VC-FISABIO-246/2020\|EPI_ISL_436341\|2 | ···················· | ···················· | ···················· | ···················· | ·NNNNNN············· | ···················· | 1 |
| B | 875 | hCoV-19/Spain/VC-FISABIO-258/2020\|EPI_ISL_436353\|2 | ···················· | ···················· | ···················· | ···················· | ···················· | NNNNNNNNNNNNNNN····· | 1 |
| B | 876 | hCoV-19/Spain/VC-FISABIO-260/2020\|EPI_ISL_436355\|2 | ···················· | ···················· | ···················· | ···················· | NNNNNNN············· | N·············N····· | 1 |
| B | 976 | hCoV-19/Thailand/Bangkok-0076/2020\|EPI_ISL_447017\| | ···················· | ···················· | ···················· | ···················· | NNNNNNNNNNNNNNNNNNNN | NNNNNNNNNNNNNNNNNNNN | 1 |
| B | 1090 | hCoV-19/Spain/VC-FISABIO-208/2020\|EPI_ISL_436303\|2 | ···················· | ···················· | ···················· | ···················· | NNNNNNN············· | ···················· | 1 |
| B | 1093 | hCoV-19/Spain/VC-FISABIO-215/2020\|EPI_ISL_436310\|2 | ···················· | ···················· | ···················· | ···················· | ··NNNNN············· | NNNNNNN············· | 1 |
| B | 1098 | hCoV-19/Spain/VC-FISABIO-269/2020\|EPI_ISL_436364\|2 | ···················· | ···················· | ···················· | ···················· | NNNNNNNNNNNNNNNNNNNN | NNNNNNNNNNNNNNNNNNNN | 1 |
| B | 1099 | hCoV-19/Spain/VC-FISABIO-271/2020\|EPI_ISL_436366\|2 | ···················· | ···················· | ···················· | ···················· | NNNNNNNNNNNNNNNNNNNN | ···················· | 1 |
| B | 1100 | hCoV-19/Spain/VC-FISABIO-272/2020\|EPI_ISL_436367\|2 | ···················· | ···················· | ···················· | ···················· | NNNNNNNNNNNNNNNNNNNN | NNNNNNNNNNNNNNNNNNNN | 1 |
| B | 1101 | hCoV-19/Spain/VC-FISABIO-273/2020\|EPI_ISL_436368\|2 | ···················· | ···················· | ···················· | ···················· | ··NN················ | NNNNN··············· | 1 |
| B | 1104 | hCoV-19/Spain/VC-FISABIO-278/2020\|EPI_ISL_436373\|2 | ···················· | ···················· | ···················· | ···················· | NNNNNNNNNNNNNNNNNNNN | ···················· | 1 |
| B | 1105 | hCoV-19/Spain/VC-FISABIO-279/2020\|EPI_ISL_436374\|2 | ···················· | ···················· | ···················· | ···················· | NNNNNNNNNNNNNNNNNNNN | NNNNNNNNNNNNNNNNNNNN | 1 |
| B | 1106 | hCoV-19/Spain/VC-FISABIO-281/2020\|EPI_ISL_436376\|2 | ···················· | ···················· | ···················· | ···················· | NNNNNNNNNNNNNNNNNNNN | NNNNNNNNNNNNNNN····· | 1 |
| B | 1108 | hCoV-19/Spain/VC-FISABIO-283/2020\|EPI_ISL_436378\|2 | ···················· | ···················· | ···················· | ···················· | NNNNNNNNNNNNNNNNNNNN | NNNNNNNNNNNNNNN····· | 1 |
| B | 1178 | hCoV-19/Scotland/CVR2501/2020\|EPI_ISL_438917\|2020- | ···················· | ···················· | ···················· | ···················· | NNNNNNNNNNNNNNNNNNNN | NNNNNNNNNNNNNNNNNNNN | 1 |
| B | 1184 | hCoV-19/Scotland/CVR257/2020\|EPI_ISL_438928\|2020-0 | ···················· | ···················· | ···················· | ···················· | NNNNNNNNNNNNNNNNNNNN | ···················· | 1 |
| B | 1193 | hCoV-19/Japan/Donner3/2020\|EPI_ISL_438948\|2020-04- | ···················· | ···················· | ···················· | ···················· | NNNNNNNNNN·········· | ···················· | 1 |
| B | 1194 | hCoV-19/Japan/Donner4/2020\|EPI_ISL_438949\|2020-04- | ···················· | ···················· | ···················· | ···················· | NNNNNNNNNNNNNNNNNNNN | NNNNNNNNNNNNNNNNNNNN | 1 |
| B | 1196 | hCoV-19/Japan/Donner7/2020\|EPI_ISL_438952\|2020-04- | ···················· | ···················· | ···················· | ···················· | NNNNNNNNNNN········· | ···················· | 1 |
| B | 1198 | hCoV-19/Japan/Donner9/2020\|EPI_ISL_438954\|2020-04- | ···················· | ···················· | ···················· | ···················· | NNNNNNNNNNNNNNNNNNNN | NNNNNNNNNNNNNNNNNNNN | 1 |
| B | 1199 | hCoV-19/Japan/Donner11/2020\|EPI_ISL_438955\|2020-04 | ···················· | ···················· | ···················· | ···················· | NNNNNNNNNNNNNNNNNNNN | NNNNNNNNNNNNNNNNNNNN | 1 |
| B | 1201 | hCoV-19/Japan/Donner14/2020\|EPI_ISL_438958\|2020-04 | ···················· | ···················· | ···················· | ···················· | NNNNNNNNNN·········· | ···················· | 1 |
| B | 1202 | hCoV-19/Japan/Donner15/2020\|EPI_ISL_438959\|2020-04 | ···················· | ···················· | ···················· | ···················· | NNNNNNNNNNNNNNNNNNNN | NNNNNNNNNNNNNNNNNNNN | 1 |
| B | 1207 | hCoV-19/Japan/Donner25/2020\|EPI_ISL_438969\|2020-04 | ···················· | ···················· | ···················· | ···················· | NNNNNNNNNNNNNNNNNNNN | NNNNNNNNNNNNNNNNNNNN | 1 |
| B | 1261 | hCoV-19/USA/IL-NM08/2020\|EPI_ISL_444521\|2020-03-17 | ···················· | ···················· | ···················· | ···················· | NNNNNNNNNNNNNNNNNNNN | NNNNNNNNNNNNNNNNNNNN | 1 |
| B | 1262 | hCoV-19/USA/IL-NM09/2020\|EPI_ISL_444522\|2020-03-17 | ···················· | ···················· | ···················· | ···················· | NNNNNNNNNNNNNNNNNNNN | NNNNNNNNNNNNNNNNNNNN | 1 |
| B | 1263 | hCoV-19/USA/IL-NM010/2020\|EPI_ISL_444523\|2020-03-1 | ···················· | ···················· | ···················· | ···················· | N·······N··N··N····· | NN····NN············ | 1 |
| B | 1264 | hCoV-19/USA/IL-NM012/2020\|EPI_ISL_444524\|2020-03-1 | ···················· | ···················· | ···················· | ···················· | NNNNNNNNNNNNNNNNNNNN | NNNNNNNNNNNNNNNNNNNN | 1 |
| B | 1266 | hCoV-19/USA/IL-NM020/2020\|EPI_ISL_444530\|2020-03-1 | ···················· | ···················· | ···················· | ···················· | NNNNNNNNNNNNNNNNNNNN | NNNNNNNNNNNNNNNNNNNN | 1 |
| B | 1267 | hCoV-19/USA/IL-NM021/2020\|EPI_ISL_444531\|2020-03-1 | ···················· | ···················· | ···················· | ···················· | NNNNNNNNNNNNNNNNNNNN | NNNNNNNNNNNNNNNNNNNN | 1 |
| B | 1280 | hCoV-19/Thailand/Bangkok-0085/2020\|EPI_ISL_447026\| | ···················· | ···················· | ···················· | ···················· | NNNNNNNNNNNNNNNNNNNN | NNNNNNNNNNNNNNNNNNNN | 1 |
| B | 1313 | hCoV-19/England/CAMB-79B10/2020\|EPI_ISL_433825\|202 | ···················· | ···················· | ···················· | ···················· | NNNNNNNNNNNNNNNNNNNN | NNNNNNNNNNNNNNNNNNNN | 1 |
| B | 1491 | hCoV-19/USA/IL-NM017/2020\|EPI_ISL_444527\|2020-03-1 | ···················· | ···················· | ···················· | ···················· | NNNNNNNNNNNNNNNNNNNN | NNNNNNNNNNNNNNNNNNNN | 1 |
| B | 1492 | hCoV-19/USA/IL-NM018/2020\|EPI_ISL_444528\|2020-03-1 | ···················· | ···················· | ···················· | ···················· | NNNNNNNNNNNNNNNNNNNN | NNNNNNNNNNNNNNNNNNNN | 1 |
| B | 1493 | hCoV-19/USA/IL-NM024/2020\|EPI_ISL_444534\|2020-03-1 | ···················· | ···················· | ···················· | ···················· | NNNNNNNNNNNNNNNNNNNN | NNNNNNNNNNNNNNNNNNNN | 1 |
| B | 1494 | hCoV-19/USA/IL-NM025/2020\|EPI_ISL_444535\|2020-03-1 | ···················· | ···················· | ···················· | ···················· | NNNNNNNNNNNNNNNNNNNN | NNNNNNNNNNNNNNNNNNNN | 1 |
| B | 1495 | hCoV-19/USA/IL-NM027/2020\|EPI_ISL_444537\|2020-03-1 | ···················· | ···················· | ···················· | ···················· | NNNNNNNNNNNNNNNNNNNN | NNNNNNNNNNNNNNNNNNNN | 1 |
| B | 1496 | hCoV-19/USA/IL-NM029/2020\|EPI_ISL_444538\|2020-03-1 | ···················· | ···················· | ···················· | ···················· | NNNNNNNNNNNNNNNNNNNN | NNNNNNNNNNNNNNNNNNNN | 1 |
| B | 1497 | hCoV-19/USA/IL-NM032/2020\|EPI_ISL_444540\|2020-03-1 | ···················· | ···················· | ···················· | ···················· | NNNNNNNNNNNNNNNNNNNN | NNNNNNNNNNNNNNNNNNNN | 1 |
| B | 1498 | hCoV-19/USA/IL-NM045/2020\|EPI_ISL_444544\|2020-03-1 | ···················· | ···················· | ···················· | ···················· | NNNNNNNNNNNNNNNNNNNN | NNNNNNNNNNNNNNNNNNNN | 1 |
| B | 1499 | hCoV-19/USA/IL-NM047/2020\|EPI_ISL_444545\|2020-03-1 | ···················· | ···················· | ···················· | ···················· | NNNNNNNNNNNNNNNNNNNN | NNNNNNNNNNNNNNNNNNNN | 1 |
| B | 1500 | hCoV-19/USA/IL-NM049/2020\|EPI_ISL_444546\|2020-03-1 | ···················· | ···················· | ···················· | ···················· | NNNNNNNNNNNNNNNNNNNN | NNNNNNNNNNNNNNNNNNNN | 1 |
| B | 1501 | hCoV-19/USA/IL-NM050/2020\|EPI_ISL_444547\|2020-03-1 | ···················· | ···················· | ···················· | ···················· | NNNNNNNNNNNNNNNNNNNN | NNNNNNNNNNNNNNNNNNNN | 1 |
| B | 1502 | hCoV-19/USA/IL-NM053/2020\|EPI_ISL_444549\|2020-03-1 | ···················· | ···················· | ···················· | ···················· | NNNNNNNNNNNNNNNNNNNN | NNNNNNNNNNNNNNNNNNNN | 1 |
| B | 1505 | hCoV-19/USA/IL-NM057/2020\|EPI_ISL_444553\|2020-03-1 | ···················· | ···················· | ···················· | ···················· | NNNNNNNNNNNNNNNNNNNN | NNNNNNNNNNNNNNNNNNNN | 1 |
| B | 1508 | hCoV-19/USA/IL-NM062/2020\|EPI_ISL_444558\|2020-03-1 | ···················· | ···················· | ···················· | ···················· | NNNNNNNNNNNNNNNNNNNN | NNNNNNNNNNNNNNNNNNNN | 1 |
| B | 1510 | hCoV-19/USA/IL-NM066/2020\|EPI_ISL_444561\|2020-03-1 | ···················· | ···················· | ···················· | ···················· | NNNNNNNNNNNNNNNNNNNN | NNNNNNNNNNNNNNNNNNNN | 1 |
| B | 1511 | hCoV-19/USA/IL-NM067/2020\|EPI_ISL_444562\|2020-03-1 | ···················· | ···················· | ···················· | ···················· | NNNNNNNNNNNNNNNNNNNN | NNNNNNNNNNNNNNNNNNNN | 1 |
| B | 1512 | hCoV-19/USA/IL-NM069/2020\|EPI_ISL_444564\|2020-03-1 | ···················· | ···················· | ···················· | ···················· | NNNNNNNNNNNNNNNNNNNN | NNNNNNNNNNNNNNNNNNNN | 1 |
| B | 1519 | hCoV-19/USA/IL-NM081/2020\|EPI_ISL_444572\|2020-03-1 | ···················· | ···················· | ···················· | ···················· | NNNNNNNNNNNNNNNNNNNN | NNNNNNNNNNNNNNNNNNNN | 1 |
| B | 1581 | hCoV-19/DRC/2864/2020\|EPI_ISL_437351\|2020-04-16 | ···················· | ···················· | ···················· | ···················· | ···················· | ·NNNNNNN···NNN······ | 1 |
| B | 1750 | hCoV-19/Singapore/99/2020\|EPI_ISL_443191\|2020-04-1 | ···················· | ···················· | ···················· | ···················· | NNNNNNNNNNNNNNNNNNNN | ···················· | 1 |
| B | 1865 | hCoV-19/Spain/VC-FISABIO-102/2020\|EPI_ISL_436197\|2 | ···················· | ···················· | ···················· | ···················· | ··NNNNN············· | NNNN················ | 1 |
| B | 2049 | hCoV-19/Australia/VIC1086/2020\|EPI_ISL_430597\|2020 | ···················· | ···················· | ···················· | ···················· | NNNNNNN············· | ···················· | 1 |
| B | 2060 | hCoV-19/Australia/VIC1247/2020\|EPI_ISL_436113\|2020 | ···················· | ···················· | ···················· | ···················· | NNNNNNNNNNNNNNNNNNNN | NNNNNNNNNNNNNNNNNNNN | 1 |
| B | 2061 | hCoV-19/Australia/VIC1248/2020\|EPI_ISL_436114\|2020 | ···················· | ···················· | ···················· | ···················· | NNNNNNNNNNNNNNNNNNNN | NNNNNNNNNNNNNNNNNNNN | 1 |
| B | 2062 | hCoV-19/Australia/VIC1255/2020\|EPI_ISL_436119\|2020 | ···················· | ···················· | ···················· | ···················· | NNNNNNNNNNNNNNNNNNNN | NNNNNNNNNNNNNNNNNNNN | 1 |
| B | 2063 | hCoV-19/Australia/VIC1256/2020\|EPI_ISL_436120\|2020 | ···················· | ···················· | ···················· | ···················· | NNNNNNNNNNNNNNNNNNNN | NNNNNNNNNNNNNNNNNNNN | 1 |
| B | 2064 | hCoV-19/Australia/VIC1267/2020\|EPI_ISL_436126\|2020 | ···················· | ···················· | ···················· | ···················· | NNNNNNNNNNNNNNNNNNNN | NNNNNNNNNNNNNNNNNNNN | 1 |
| B | 2065 | hCoV-19/Australia/VIC1273/2020\|EPI_ISL_436131\|2020 | ···················· | ···················· | ···················· | ···················· | NNNNNNNNNNNNNNNNNNNN | NNNNNNNNNNNNNNNNNNNN | 1 |
| B | 2098 | hCoV-19/env/USA/MT-BWRF-0327/2020\|EPI_ISL_437434\|2 | ···················· | ···A················ | ···················· | ···················· | ···CC·NNNNNNNNNNNNNN | NNNNNNNNNNNNNNNNNNNN | 1 |
| B | 2232 | hCoV-19/USA/MN-UMGC-5/2020\|EPI_ISL_443256\|2020-04- | ···················· | ···················· | ···················· | ···················· | NNNNNNNNNNNNNNNNNNNN | NNNNNNNNNNNNNNNNNNNN | 1 |
| B | 2279 | hCoV-19/USA/IL-NM091/2020\|EPI_ISL_444581\|2020-03-1 | ···················· | ···················· | ···················· | ···················· | NNNNNNNNNNNNNNNNNNNN | NNNNNNNNNNNNNNNNNNNN | 1 |
| B | 2305 | hCoV-19/Australia/VIC1210/2020\|EPI_ISL_430500\|2020 | ···················· | ···················· | ···················· | ···················· | NNNNNNNNNNNNNNNNNNNN | NNNNNNNNNNNNNNNNNNNN | 1 |
| B | 2306 | hCoV-19/Australia/VIC1211/2020\|EPI_ISL_430501\|2020 | ···················· | ···················· | ···················· | ···················· | NNNNNNNNNNNNNNNNNNNN | NNNNNNNNNNNNNNNNNNNN | 1 |
| B | 2307 | hCoV-19/Australia/VIC1214/2020\|EPI_ISL_430505\|2020 | ···················· | ···················· | ···················· | ···················· | NNNNNNNNNNNNNNNNNNNN | NNNNNNNNNNNNNNNNNNNN | 1 |
| B | 2308 | hCoV-19/Australia/VIC1198/2020\|EPI_ISL_430507\|2020 | ···················· | ···················· | ···················· | ···················· | NNNNNNNNNNNNNNNNNNNN | NNNNNNNNNNNNNNNNNNNN | 1 |
| B | 2309 | hCoV-19/Australia/VIC1235/2020\|EPI_ISL_430512\|2020 | ···················· | ···················· | ···················· | ···················· | NNNNNNNNNNNNNNNNNNNN | NNNNNNNNNNNNNNNNNNNN | 1 |

| | | | | | | | | | |
|---|---|---|---|---|---|---|---|---|---|
| B | 2310 | hCoV-19/Australia/VIC1222/2020\|EPI_ISL_430519\|2020 | ···················· | ···················· | ···················· | ···················· | NNNNNNNNNNNNNNNNNNNN | NNNNNNNNNNNNNNNNNNNN | 1 |
| B | 2311 | hCoV-19/Australia/VIC1189/2020\|EPI_ISL_430521\|2020 | ···················· | ···················· | ···················· | ···················· | NNNNNNNNNNNNNNNNNNNN | NNNNNNNNNNNNNNNNNNNN | 1 |
| B | 2312 | hCoV-19/Australia/VIC1224/2020\|EPI_ISL_430522\|2020 | ···················· | ···················· | ···················· | ···················· | NNNNNNNNNNNNNNNNNNNN | NNNNNNNNNNNNNNNNNNNN | 1 |
| B | 2313 | hCoV-19/Australia/VIC1199/2020\|EPI_ISL_430523\|2020 | ···················· | ···················· | ···················· | ···················· | NNNNNNNNNNNNNNNNNNNN | NNNNNNNNNNNNNNNNNNNN | 1 |
| B | 2314 | hCoV-19/Australia/VIC1200/2020\|EPI_ISL_430527\|2020 | ···················· | ···················· | ···················· | ···················· | NNNNNNNNNNNNNNNNNNNN | NNNNNNNNNNNNNNNNNNNN | 1 |
| B | 2315 | hCoV-19/Australia/VIC1192/2020\|EPI_ISL_430528\|2020 | ···················· | ···················· | ···················· | ···················· | NNNNNNNNNNNNNNNNNNNN | NNNNNNNNNNNNNNNNNNNN | 1 |
| B | 2316 | hCoV-19/Australia/VIC1023/2020\|EPI_ISL_430530\|2020 | ···················· | ···················· | ···················· | ···················· | NNNNNNNNNNNNNNNNNNNN | NNNNNNNNNNNNNNNNNNNN | 1 |
| B | 2318 | hCoV-19/Australia/VIC1025/2020\|EPI_ISL_430532\|2020 | ···················· | ···················· | ···················· | ···················· | NNNNNNNNNNNNNNNNNNNN | NNNNNNNNN··········· | 1 |
| B | 2320 | hCoV-19/Australia/VIC1039/2020\|EPI_ISL_430535\|2020 | ···················· | ···················· | ···················· | ···················· | NNNNNNNNNNNNNNNNNNNN | ···················· | 1 |
| B | 2328 | hCoV-19/Australia/VIC1011/2020\|EPI_ISL_430547\|2020 | ···················· | ···················· | ···················· | ···················· | NNNNNNN············· | ···················· | 1 |
| B | 2329 | hCoV-19/Australia/VIC1012/2020\|EPI_ISL_430548\|2020 | ···················· | ···················· | ···················· | ···················· | NNNNNNNNNNNNNNNN···· | NNNNNNNNNN·········· | 1 |
| B | 2332 | hCoV-19/Australia/VIC984/2020\|EPI_ISL_430552\|2020- | ···················· | ···················· | ···················· | ···················· | NNNNNNNNNNNNNNNNNNNN | NNNNNNNNNNNNNNNNNNNN | 1 |
| B | 2333 | hCoV-19/Australia/VIC1013/2020\|EPI_ISL_430553\|2020 | ···················· | ···················· | ···················· | ···················· | NNNNNNNNNNNNNNNNNNNN | NNNNNNNNNNNNNNN····· | 1 |
| B | 2336 | hCoV-19/Australia/VIC1015/2020\|EPI_ISL_430559\|2020 | ···················· | ···················· | ···················· | ···················· | NNNNNNNNNNNNNNNNNNNN | ···················· | 1 |
| B | 2337 | hCoV-19/Australia/VIC1047/2020\|EPI_ISL_430561\|2020 | ···················· | ···················· | ···················· | ···················· | NNNNNNNNNNNNNNNNNNNN | NNNNNNNNNNNNNNNNNNNN | 1 |
| B | 2338 | hCoV-19/Australia/VIC1048/2020\|EPI_ISL_430562\|2020 | ···················· | ···················· | ···················· | ···················· | NNNNNNNNNNNNNNNNNNNN | NNNNNNNNNNNNN······· | 1 |
| B | 2339 | hCoV-19/Australia/VIC1119/2020\|EPI_ISL_430567\|2020 | ···················· | ···················· | ···················· | ···················· | NNNNNNNNNNNNNNNNNNNN | ···················· | 1 |
| B | 2340 | hCoV-19/Australia/VIC1110/2020\|EPI_ISL_430569\|2020 | ···················· | ···················· | ···················· | ···················· | NNNNNNNNNNNNNNNNNNNN | ······M············· | 1 |
| B | 2342 | hCoV-19/Australia/VIC1087/2020\|EPI_ISL_430574\|2020 | ···················· | ···················· | ···················· | ···················· | NNNNNNNNNNNNNNNNNNNN | ···················· | 1 |
| B | 2343 | hCoV-19/Australia/VIC1113/2020\|EPI_ISL_430575\|2020 | ···················· | ···················· | ···················· | ···················· | NNNNNNNNNNNNNNNNNNNN | NNNNNNNNNNNNNNNNNNNN | 1 |
| B | 2344 | hCoV-19/Australia/VIC1114/2020\|EPI_ISL_430576\|2020 | ···················· | ···················· | ···················· | ···················· | NNNNNNNNNNNNNNNNNNNN | NNNNNNNNNNNNNNNNNNNN | 1 |
| B | 2348 | hCoV-19/Australia/VIC1081/2020\|EPI_ISL_430580\|2020 | ···················· | ···················· | ···················· | ···················· | NNNNNNNNNNNNNNNNNNNN | NNNN················ | 1 |
| B | 2349 | hCoV-19/Australia/VIC1071/2020\|EPI_ISL_430581\|2020 | ···················· | ···················· | ···················· | ···················· | NNNNNNN············· | NNNNNNNNN··········· | 1 |
| B | 2350 | hCoV-19/Australia/VIC1094/2020\|EPI_ISL_430583\|2020 | ···················· | ···················· | ···················· | ···················· | NNNNNNN············· | ···················· | 1 |
| B | 2351 | hCoV-19/Australia/VIC1103/2020\|EPI_ISL_430584\|2020 | ···················· | ···················· | ···················· | ···················· | NNNNNNN············· | ···················· | 1 |
| B | 2352 | hCoV-19/Australia/VIC1104/2020\|EPI_ISL_430585\|2020 | ···················· | ···················· | ···················· | ···················· | NNNNNNNNNNNNNNNNNNNN | NNNNNNNNNNNNNNNNNNNN | 1 |
| B | 2353 | hCoV-19/Australia/VIC1085/2020\|EPI_ISL_430587\|2020 | ···················· | ···················· | ···················· | ···················· | NNNNNNNNNNNNNNNNNNNN | NNNNNNNNNNNNNNNNNNNN | 1 |
| B | 2354 | hCoV-19/Australia/VIC1107/2020\|EPI_ISL_430590\|2020 | ···················· | ···················· | ···················· | ···················· | NNNNNNNNNNNNNNNNNNNN | NNNNNNNNNNNNNNNNNNNN | 1 |
| B | 2355 | hCoV-19/Australia/VIC1133/2020\|EPI_ISL_430596\|2020 | ···················· | ···················· | ···················· | ···················· | NNNNNNNNNNNNNNNNNNNN | NNNNNNNNNNNNNNNNNNNN | 1 |
| B | 2372 | hCoV-19/Australia/VIC1230/2020\|EPI_ISL_436688\|2020 | ···················· | ···················· | ···················· | ···················· | NNNNNNNNNNNNNNNNNNNN | NNNNNNNNNNNNNNNNNNNN | 1 |
| B | 2373 | hCoV-19/Australia/VIC550/2020\|EPI_ISL_436689\|2020- | ···················· | ···················· | ···················· | ···················· | NNNNNNNNNNNNNNNNNNNN | NNNNNNNNNNNNNNNNNNNN | 1 |
| B | 2817 | hCoV-19/Australia/VIC1139/2020\|EPI_ISL_430610\|2020 | ···················· | ···················· | ···················· | ···················· | NNNNNNNNNNNNNNNNNNNN | NNNNNNNNNNNNNNNNNNNN | 1 |
| B | 2818 | hCoV-19/Australia/VIC1143/2020\|EPI_ISL_430614\|2020 | ···················· | ···················· | ···················· | ···················· | NNNNNNNNNNNNNNNNNNNN | NNNNNNNNNNNNNNNNNNNN | 1 |
| B | 2819 | hCoV-19/Australia/VIC1161/2020\|EPI_ISL_430615\|2020 | ···················· | ···················· | ···················· | ···················· | NNNNNNN············· | ···················· | 1 |
| B | 2820 | hCoV-19/Australia/VIC1162/2020\|EPI_ISL_430616\|2020 | ···················· | ···················· | ···················· | ···················· | NNNNNNNNNNNNNNNNNNNN | NNNNNNNNNNNNNNNNNNNN | 1 |
| B | 2821 | hCoV-19/Australia/VIC1109/2020\|EPI_ISL_430618\|2020 | ···················· | ···················· | ···················· | ···················· | NNNNNNNNNNNNNNNNNNNN | NNN················· | 1 |
| B | 2822 | hCoV-19/Australia/VIC1144/2020\|EPI_ISL_430619\|2020 | ···················· | ···················· | ···················· | ···················· | ···················· | NNNN················ | 1 |
| B | 2823 | hCoV-19/Australia/NT29/2020\|EPI_ISL_430632\|2020 | ···················· | ···················· | ···················· | ···················· | NNNNNNNNNNNNNNNNNNNN | NNNNNNNNNNNNNNNNNNNN | 1 |
| B | 2824 | hCoV-19/Australia/NT30/2020\|EPI_ISL_430633\|2020 | ···················· | ···················· | ···················· | ···················· | NNNNNNN············· | NNNN················ | 1 |
| B | 2825 | hCoV-19/Australia/NT31/2020\|EPI_ISL_430635\|2020 | ···················· | ···················· | ···················· | ···················· | NNNNNNNNNNNNNNNNNNNN | NNNNNNNNNNNNNNNNNNNN | 1 |
| B | 2826 | hCoV-19/Australia/NT32/2020\|EPI_ISL_430636\|2020 | ···················· | ···················· | ···················· | ···················· | NNNNNNNNNNNNNNNNNNNN | NNNNNNNNNNNNNNNNNNNN | 1 |
| B | 2827 | hCoV-19/Australia/VIC1355/2020\|EPI_ISL_430638\|2020 | ···················· | ···················· | ···················· | ···················· | NNNNNNNNNNNNNNNNNNNN | NNNNNNNNNNNNNNNNNNNN | 1 |
| B | 2829 | hCoV-19/Australia/VIC946/2020\|EPI_ISL_430640\|2020- | ···················· | ···················· | ···················· | ···················· | NNNNNNNNNNNNNNNNNNNN | NNNNNNNNNNNNNNNNNNNN | 1 |
| B | 2830 | hCoV-19/Australia/VIC955/2020\|EPI_ISL_430643\|2020- | ···················· | ···················· | ···················· | ···················· | NNNNNNNNNNNNNNNNN··· | ··········NNNNN····· | 1 |
| B | 2831 | hCoV-19/Australia/VIC953/2020\|EPI_ISL_430645\|2020- | ···················· | ···················· | ···················· | ···················· | NNNNNNNNNNNNNNNNNNNN | NNNNNNNNNNNNNNNNNNNN | 1 |
| B | 2833 | hCoV-19/Australia/VIC948/2020\|EPI_ISL_430647\|2020- | ···················· | ···················· | ···················· | ···················· | NNNNNNNNNNNNNNNNNNNN | NNNNNNNNNNNNNNNNNNNN | 1 |
| B | 2834 | hCoV-19/Australia/VIC957/2020\|EPI_ISL_430649\|2020- | ···················· | ···················· | ···················· | ···················· | NNNNNNNNNNNNNNNNNNNN | NNNNNNNNNNNNNNNNNNNN | 1 |
| B | 2835 | hCoV-19/Australia/VIC968/2020\|EPI_ISL_430651\|2020- | ···················· | ···················· | ···················· | ···················· | NNNNNNNNNNNNNNNNNNNN | NNNNNNNNNNNNNNNNNNNN | 1 |
| B | 2982 | hCoV-19/Northern Ireland/NIRE-101F01/2020\|EPI_ISL_ | ···················· | ···················· | ···················· | ···················· | NNNNNNNNNNNNNNNNNNNN | NNNNNNNNNNNNNNN····· | 1 |
| B | 2984 | hCoV-19/Northern Ireland/NIRE-101F5C/2020\|EPI_ISL_ | ···················· | ···················· | ···················· | ···················· | ······RNNNNNNNNNNNNN | ···················· | 1 |
| B | 2987 | hCoV-19/Northern Ireland/NIRE-101F89/2020\|EPI_ISL_ | ···················· | ···················· | ···················· | ···················· | NNNNNNNNNNNNNNNNNNNN | NNNNNNNNNNNNNNNNNNNN | 1 |
| B | 2990 | hCoV-19/Northern Ireland/NIRE-101FF2/2020\|EPI_ISL_ | ···················· | ···················· | ···················· | ···················· | NNNNNNNNNNNNNNNNNNNN | NNNNNNNNNNNNNNNNNNNN | 1 |
| B | 2991 | hCoV-19/Northern Ireland/NIRE-10200D/2020\|EPI_ISL_ | ···················· | ···················· | ···················· | ···················· | NNNNNNNNNNNNNNNNNNNN | NNNNNNNNNNNNNNNNNNNN | 1 |
| B | 2993 | hCoV-19/Northern Ireland/NIRE-10202B/2020\|EPI_ISL_ | ···················· | ···················· | ···················· | ···················· | NNNNNNNNNNNNNNNNNNNN | ···················· | 1 |
| B | 3000 | hCoV-19/Northern Ireland/NIRE-1020A3/2020\|EPI_ISL_ | ···················· | ···················· | ···················· | ···················· | NNNNNNNNNNNNNNNNNNNN | ···················· | 1 |
| B | 3002 | hCoV-19/Northern Ireland/NIRE-1020D0/2020\|EPI_ISL_ | ···················· | ···················· | ···················· | ···················· | ·NNNNNNNNN·········· | ···················· | 1 |
| B | 3003 | hCoV-19/Northern Ireland/NIRE-1020EF/2020\|EPI_ISL_ | ···················· | ···················· | ···················· | ···················· | NNNNNNNNNNNNNNNNNNNN | ···················· | 1 |
| B | 3005 | hCoV-19/Northern Ireland/NIRE-10210A/2020\|EPI_ISL_ | ···················· | ···················· | ···················· | ···················· | NNNNNNNNNNNNNNNNNNNN | ···················· | 1 |
| B | 3007 | hCoV-19/Northern Ireland/NIRE-1022CB/2020\|EPI_ISL_ | ···················· | ···················· | ···················· | ···················· | NNNNNNNNNNNNNNNNNNNN | ···················· | 1 |
| B | 3073 | hCoV-19/Australia/VIC1174/2020\|EPI_ISL_430623\|2020 | ···················· | ···················· | ···················· | ···················· | NNNNNNNNN··········· | NNNN················ | 1 |
| B | 3074 | hCoV-19/Australia/VIC966/2020\|EPI_ISL_430653\|2020- | ···················· | ···················· | ···················· | ···················· | NNNNNNN············· | ···················· | 1 |
| B | 3075 | hCoV-19/Australia/VIC961/2020\|EPI_ISL_430654\|2020- | ···················· | ···················· | ···················· | ···················· | NNNNNNNNNNNNNNNNNNNN | NNNNNNNNN··NNNNNNNNN | 1 |
| B | 3077 | hCoV-19/Australia/VIC975/2020\|EPI_ISL_430658\|2020- | ···················· | ···················· | ···················· | ···················· | NNNNNNNNNNNNNNNNNNNN | NNNNNNNNNNNNNNNNNNNN | 1 |
| B | 3078 | hCoV-19/Australia/VIC974/2020\|EPI_ISL_430659\|2020- | ···················· | ···················· | ···················· | ···················· | NNNNNNNNNNNNNNNNNNNN | NNNNNNNNNNNNNNNNNNNN | 1 |
| B | 3079 | hCoV-19/Australia/VIC971/2020\|EPI_ISL_430661\|2020- | ···················· | ···················· | ···················· | ···················· | NNNNNNNNNNNNNNNNNNNN | NNNNNNNNNNNNNNNNNNNN | 1 |
| B | 3080 | hCoV-19/Australia/VIC959/2020\|EPI_ISL_430663\|2020- | ···················· | ···················· | ···················· | ···················· | NNNNNNNNNNNNNNNNNNNN | ···················· | 1 |
| B | 3081 | hCoV-19/Australia/VIC1056/2020\|EPI_ISL_430669\|2020 | ···················· | ···················· | ···················· | ···················· | NNNNNNN············· | ··············N····· | 1 |
| B | 3083 | hCoV-19/Australia/VIC1089/2020\|EPI_ISL_430673\|2020 | ···················· | ···················· | ···················· | ···················· | NNNNNNNNNNNNNNNNNNNN | NNNNNNNNNNNNNNNNNNNN | 1 |
| B | 3085 | hCoV-19/Australia/VIC1281/2020\|EPI_ISL_430676\|2020 | ···················· | ···················· | ···················· | ···················· | ··NNNNN············· | ···················· | 1 |
| B | 3086 | hCoV-19/Australia/VIC1300/2020\|EPI_ISL_430677\|2020 | ···················· | ···················· | ···················· | ···················· | ···················· | NNNNNNNNN··········· | 1 |
| B | 3087 | hCoV-19/Australia/VIC1279/2020\|EPI_ISL_430678\|2020 | ···················· | ···················· | ···················· | ···················· | ···················· | NNNNNN·············· | 1 |
| B | 3094 | hCoV-19/Australia/VIC1340/2020\|EPI_ISL_430686\|2020 | ···················· | ···················· | ···················· | ···················· | NNNNNNNNNNNNNNNNNNNN | ···················· | 1 |
| B | 3098 | hCoV-19/Australia/VIC1317/2020\|EPI_ISL_430691\|2020 | ·········N·········· | ···················· | ···················· | ···················· | NNNNNNN············· | ···················· | 1 |
| B | 3101 | hCoV-19/Australia/VIC1328/2020\|EPI_ISL_430695\|2020 | ···················· | ···················· | ···················· | ···················· | NNNNNNN···NNNNN····· | ···················· | 1 |
| B | 3353 | hCoV-19/Australia/VIC1134/2020\|EPI_ISL_430600\|2020 | ···················· | ···················· | ···················· | ···················· | NNNNNNNNNNNNNNNNNNNN | NNNNNNNNNNNNNNNNNNNN | 1 |
| B | 3355 | hCoV-19/Australia/VIC1158/2020\|EPI_ISL_430602\|2020 | ···················· | ···················· | ···················· | ···················· | ···················· | NNNN················ | 1 |
| B | 3357 | hCoV-19/Australia/VIC1160/2020\|EPI_ISL_430605\|2020 | ···················· | ···················· | ···················· | ···················· | NNNNNNNNNNNNNNNNNNNN | NNNNNNNNNNNNNNNNNNNN | 1 |
| B | 3358 | hCoV-19/Australia/VIC1137/2020\|EPI_ISL_430607\|2020 | ···················· | ···················· | ···················· | ···················· | NNNNNNNNNNNNNNNNNNNN | NNNNNNNNNNNNNNNNNNNN | 1 |
| B | 3359 | hCoV-19/Australia/VIC1138/2020\|EPI_ISL_430609\|2020 | ···················· | ···················· | ···················· | ···················· | NNNNNNNNNNNNNNNNNNNN | NNNNNNNNNNNNNNNNNNNN | 1 |
| B | 4118 | hCoV-19/Australia/VIC935/2020\|EPI_ISL_430471\|2020- | ···················· | ···················· | ···················· | ···················· | NNNNNNNNNNNNNNNNNNNN | NNNNNNNNNNNNNNNNNNNN | 1 |

```
B 4119 hCoV-19/Australia/VIC938/2020|EPI_ISL_430472|2020- ···················· ···················· ···················· ···················· ···················· NNNNNNNNNNNNNNNNNNNN 1
B 4120 hCoV-19/Australia/VIC1008/2020|EPI_ISL_430473|2020 ···················· ···················· ···················· ···················· NNNNNNNNNNNNNNNNNNNN NNNNNNNNNNNNNNNNNNNN 1
B 4121 hCoV-19/Australia/VIC991/2020|EPI_ISL_430474|2020- ···················· ···················· ···················· ···················· NNNNNNNNNNNNNNNNNNNN NNNNNNNNNNNNNNNNNNNN 1
B 4122 hCoV-19/Australia/VIC1016/2020|EPI_ISL_430476|2020 ···················· ···················· ···················· ···················· NNNNNNNNNNNNNNNNNNNN NNNNNNNNNNNNNNNNNNNN 1
B 4123 hCoV-19/Australia/VIC994/2020|EPI_ISL_430478|2020- ···················· ···················· ···················· ···················· NNNNNNNNNNNNNNNNNNNN NNNNNNNNNNNNNNNNNNNN 1
B 4124 hCoV-19/Australia/VIC995/2020|EPI_ISL_430479|2020- ···················· ···················· ···················· ···················· NNNNNNN············· ···················· 1
B 4125 hCoV-19/Australia/VIC996/2020|EPI_ISL_430480|2020- ···················· ···················· ···················· ···················· NNNNNNNNNNNNNNNNNNNN NNNN················ 1
B 4126 hCoV-19/Australia/VIC997/2020|EPI_ISL_430481|2020- ···················· ···················· ···················· ···················· NNNNNNNNNNNNNNNNNNNN NNNNNNNNNNNNNNNNNNNN 1
B 4127 hCoV-19/Australia/VIC998/2020|EPI_ISL_430482|2020- ···················· ···················· ···················· ···················· NNNNNNNNNNNNNNNNNNNN NNNNNNNNNNNNNNNNNNNN 1
B 4128 hCoV-19/Australia/VIC1000/2020|EPI_ISL_430484|2020 ···················· ···················· ···················· ···················· NNNNNNN············· NNN················· 1
B 4129 hCoV-19/Australia/VIC1002/2020|EPI_ISL_430486|2020 ···················· ···················· ···················· ···················· NNNNNNNNNNNNNNNNNNNN NNNNNNNNNNNNNNNNNNNN 1
B 4130 hCoV-19/Australia/VIC1018/2020|EPI_ISL_430490|2020 ···················· ···················· ···················· ···················· NNNNNNNNNNNNNNNNNNNN NNNNNNNNNNNNNNNNNNNN 1
B 4131 hCoV-19/Australia/VIC1208/2020|EPI_ISL_430498|2020 ···················· ···················· ···················· ···················· NNNNNNNNNNNNNNNNNNNN NNNNNNNNNNNNNNNNNNNN 1
B 4606 hCoV-19/DRC/3829/2020|EPI_ISL_447246|2020-04-24 ···················· ···················· ···················· ···················· ··N·NNN·NNNN····NNN· ···················· 1
B 4619 hCoV-19/Scotland/EDB576/2020|EPI_ISL_433201|2020-0 ···················· ···················· ···················· ···················· NNNNNNNNNNNNNNNNNNNN NNNNNNNNNNNNNNNNNNNN 1
B 4620 hCoV-19/Scotland/EDB578/2020|EPI_ISL_433202|2020-0 ···················· ···················A ···················· ···················· NNNNNNNNNNNNNNNNNNNN NNNNNNNNNNNNNNNNNNNN 1
B 4621 hCoV-19/Scotland/EDB580/2020|EPI_ISL_433203|2020-0 ···················· ···················· ···················· ···················· NNNNNNNNNNNNNNNNNNNN NNNNNNNNNNNNNNNNNNNN 1
B 4634 hCoV-19/USA/LA-ALSR-0058-NBG/2020|EPI_ISL_434517|2 ···················· ···················· ···················· ···················· NNNNNNN············· ···················· 1
B 4635 hCoV-19/USA/LA-ALSR-0059-NBG/2020|EPI_ISL_434518|2 ···················· ···················· ···················· ···················· NNNNNNNNNNNNNNNNNNNN ···················· 1
B 4705 hCoV-19/Scotland/EDB3760/2020|EPI_ISL_439318|2020- ···················· ···················· ···················· ···················· NNNNNNNNNNNNNNNNNNNN NNNNNNNNNNNNNNNNNNNN 1
B 5140 hCoV-19/Scotland/EDB1641/2020|EPI_ISL_433100|2020- ···················· ···················· ···················· ···················· NNNNNNNNNNNNNNNNNNNN NNNNNNNNNNNNNNNNNNNN 1
B 5141 hCoV-19/Scotland/EDB1642/2020|EPI_ISL_433101|2020- ···················· ···················· ···················· ···················· ···········N··N·N··· NN·N···NN··········· 1
B 5142 hCoV-19/Scotland/EDB1644/2020|EPI_ISL_433102|2020- ···················· ·A·················· ···················· ···················· NNNNNNNNNNNNNNNNNNNN NNNNNNNNNNNNNNNNNNNN 1
B 5144 hCoV-19/Scotland/EDB1650/2020|EPI_ISL_433105|2020- ···················· ···················· ···················· ···················· NNNNNNNNNNNNNNNNNNNN NNNNNNNNNNNNNNNNNNNN 1
B 5145 hCoV-19/Scotland/EDB1652/2020|EPI_ISL_433106|2020- ···················· ···················· ···················· ···················· NNNNNNNNNNNNNNNNNNNN NNNNNNNNNNNNNNNNNNNN 1
B 5148 hCoV-19/Scotland/EDB1660/2020|EPI_ISL_433109|2020- ···················· ···················· ···················· ···················· NNNNNNNNNNNNNNNNNNNN NNNNNNNNNNNNNNNNNNNN 1
B 5150 hCoV-19/Scotland/EDB1664/2020|EPI_ISL_433111|2020- ···················· ···················· ···················· ···················· NNNNNNNNNNNNNNNNNNNN NNNNNNNNNNNNNNNNNNNN 1
B 5271 hCoV-19/Scotland/EDB3628/2020|EPI_ISL_439300|2020- ···················· ···················· ···················· ···················· NNNNNNNNNNNNNNNNNNNN NNNNNNNNNNNNNNNNNNNN 1
B 5273 hCoV-19/Scotland/EDB3633/2020|EPI_ISL_439302|2020- ···················· ···················· ···················· ···················· NNNNNNNNNNNNNNNNNNNN NNNNNNNNNNNNNNNNNNNN 1
B 5475 hCoV-19/USA/WA-S611/2020|EPI_ISL_438164|2020-04-23 ···················· ·NNNNNNNNNNNNNNNNNNN ···················· ···················· ···················· ···················· 1
B 5637 hCoV-19/Scotland/EDB590/2020|EPI_ISL_433208|2020-0 ···················· ···················· ···················· ···················· ···N····N··NN·NNN··· NN·····NN··········· 1
B 5638 hCoV-19/Scotland/EDB594/2020|EPI_ISL_433210|2020-0 ···················· ···················· ···················· ···················· NNNNNNNNNNNNNNNNNNNN NNNNNNNNNNNNNNNNNNNN 1
B 5640 hCoV-19/Scotland/EDB606/2020|EPI_ISL_433213|2020-0 ···················· ···················· ···················· ···················· NNNNNNNNNNNNNNNNNNNN NNNNNNNNNNNNNNNNNNNN 1
B 5646 hCoV-19/Scotland/EDB629/2020|EPI_ISL_433220|2020-0 ···················· ···················· ···················· ···················· NNNNNNNNNNNNNNNNNNNN NNNNNNNNNNNNNNNNNNNN 1
B 5647 hCoV-19/Scotland/EDB653/2020|EPI_ISL_433221|2020-0 ···················· ·A·················· ···················· ···················· NNNNNNNNNNNNNNNNNNNN NNNNNNNNNNNNNNNNNNNN 1
B 5771 hCoV-19/England/CAMB-7540A/2020|EPI_ISL_440407|202 ···················· ···················· ···················· ···················· NNNNNNNNNNNNNNNNNNNN NNNNNNNNNNNNNNNNNNNN 1
B 5982 hCoV-19/Scotland/EDB2137/2020|EPI_ISL_439167|2020- ···················· ···················· ···················· ···················· NNNNNNNNNNNNNNNNNNNN NNNNNNNNNNNNNNNNNNNN 1
B 5991 hCoV-19/Scotland/EDB2263/2020|EPI_ISL_439177|2020- ···················· ···················· ···················· ···················· NNNNNNNNNNNNNNNNNNNN NNNNNNNNNNNNNNNNNNNN 1
B 6064 hCoV-19/Northern Ireland/NIRE-102304/2020|EPI_ISL_ ···················· ···················· ···················· ···················· NNNNNNNNNNNNNNNNNNNN ···················· 1
B 6065 hCoV-19/Northern Ireland/NIRE-102313/2020|EPI_ISL_ ···················· ···················· ···················· ···················· ·NNNNNNNNNNNN······· ···················· 1
B 6070 hCoV-19/Northern Ireland/NIRE-10239B/2020|EPI_ISL_ ···················· ···················· ···················· ···················· NNNNNNNNNNNNNNNNNNNN ···················· 1
B 6145 hCoV-19/Australia/VIC1319/2020|EPI_ISL_430700|2020 ···················· ···················· ···················· ···················· NNNNNNN············· ···················· 1
B 6146 hCoV-19/Australia/VIC1284/2020|EPI_ISL_430701|2020 ···················· ···················· ···················· ···················· ···················· NNNNNN·············· 1
B 6149 hCoV-19/Australia/VIC1292/2020|EPI_ISL_430711|2020 ···················· ···················· ···················· ···················· NNNNNNNNNNNNNNNNNNNN NNNNNNNNNNNNNNNNNNNN 1
B 6151 hCoV-19/Australia/VIC1353/2020|EPI_ISL_430716|2020 ···················· ···················· ···················· ···················· NNNNNNNNNNNNNNNNNNNN NNNNNNNNNNNNNNNNNNNN 1
B 6526 hCoV-19/Scotland/EDB3499/2020|EPI_ISL_439272|2020- ···················· ···················· ···················· ···················· NNNNNNNNNNNNNNNNNNNN NNNNNNNNNNNNNNNNNNNN 1
B 6543 hCoV-19/Scotland/EDB3569/2020|EPI_ISL_439289|2020- ···················· ···················· ···················· ···················· NNNNNNNNNNNNNNNNNNNN NNNNNNNNNNNNNNNNNNNN 1
B 6551 hCoV-19/Scotland/EDB3603/2020|EPI_ISL_439298|2020- ···················· ·A·················· ···················· ···················· NNNNNNNNNNNNNNNNNNNN NNNNNNNNNNNNNNNNNNNN 1
B 6714 hCoV-19/Scotland/EDB1609/2020|EPI_ISL_433089|2020- ···················· ·A·················· ···················· ···················· NNNNNNNNNNNNNNNNNNNN NNNNNNNNNNNNNNNNNNNN 1
B 6715 hCoV-19/Belgium/ITM_C298/2020|EPI_ISL_434380|2020- ···················· ···················· ···················· ···················· NNNNNNNNNNNNNNNNNNNN NNNNNNNNNNNNNNNNNNNN 1
B 6716 hCoV-19/Belgium/ITM_C585/2020|EPI_ISL_434381|2020- ···················· ···················· ···················· ···················· NNNNNNNNNNNNNNNNNNNN NNNNNNNNNNNNNNNNNNNN 1
B 6717 hCoV-19/Belgium/ITM_C347/2020|EPI_ISL_434382|2020- ···················· ···················· ···················· ···················· NNNNNNNNNNNNNNNNNNNN NNNNNNNNNNNNNNNNNNNN 1
B 6718 hCoV-19/Belgium/ITM_C420/2020|EPI_ISL_434383|2020- ···················· ···················· ···················· ···················· NNNNNNNNNNNNNNNNNNNN NNNNNNNNNNNNNNNNNNNN 1
B 6719 hCoV-19/Belgium/ITM_C511/2020|EPI_ISL_434384|2020- ···················· ···················· ···················· ···················· NNNNNNNNNNNNNNNNNNNN NNNNNNNNNNNNNNNNNNNN 1
B 6765 hCoV-19/Scotland/EDB3450/2020|EPI_ISL_439265|2020- ···················· ···················A ···················· ···················· ···················· NN·N···NN··········· 1
B 6916 hCoV-19/Argentina/PAIS-A0022/2020|EPI_ISL_430813|2 ···················· ···················· ···················· ···················· NNNNNNNNNNNNNNNNNNNN ···················· 1
B 6935 hCoV-19/Scotland/EDB1610/2020|EPI_ISL_433090|2020- ···················· ·A·················· ···················· ···················· NNNNNNNNNNNNNNNNNNNN NNNNNNNNNNNNNNNNNNNN 1
B 6936 hCoV-19/Scotland/EDB1617/2020|EPI_ISL_433091|2020- ···················· ·A·················· ···················· ···················· ·N·NN····N····N····· NN·N···NN····N······ 1
B 6937 hCoV-19/Scotland/EDB1622/2020|EPI_ISL_433092|2020- ···················· ···················· ···················· ···················· NNNNNNNNNNNNNNNNNNNN NNNNNNNNNNNNNNNNNNNN 1
B 6938 hCoV-19/Scotland/EDB1629/2020|EPI_ISL_433093|2020- ···················· ···················· ···················· ···················· ··············N····· NN·····NN··········· 1
B 6939 hCoV-19/Scotland/EDB1630/2020|EPI_ISL_433094|2020- ···················· ···················· ···················· ···················· NNNNNNNNNNNNNNNNNNNN NNNNNNNNNNNNNNNNNNNN 1
B 6942 hCoV-19/Scotland/EDB1633/2020|EPI_ISL_433097|2020- ···················· ···················· ···················· ···················· NNNNNNNNNNNNNNNNNNNN NNNNNNNNNNNNNNNNNNNN 1
B 6988 hCoV-19/Belgium/ITM_C189/2020|EPI_ISL_434366|2020- ···················· ···················· ···················· ···················· NNNNNNNNNNNNNNNNNNNN NNNNNNNNNNNNNNNNNNNN 1
B 6991 hCoV-19/Belgium/ITM_C305/2020|EPI_ISL_434370|2020- ···················· ···················· ···················· ···················· NNNNNNNNNNNNNNNNNNNN NNNNNNNNNNNNNNNNNNNN 1
B 6992 hCoV-19/Belgium/ITM_C233/2020|EPI_ISL_434371|2020- ···················· ···················· ···················· ···················· NNNNNNNNNNNNNNNNNNNN NNNNNNNNNNNNNNNNNNNN 1
B 6995 hCoV-19/Belgium/ITM_C366/2020|EPI_ISL_434374|2020- ···················· ···················· ···················· ···················· NNNNNNNNNNNNNNNNNNNN NNNNNNNNNNNNNNNNNNNN 1
B 6996 hCoV-19/Belgium/ITM_C159/2020|EPI_ISL_434375|2020- ···················· ···················· ···················· ···················· NNNNNNNNNNNNNNNNNNNN NNNNNNNNNNNNNNNNNNNN 1
B 6997 hCoV-19/Belgium/ITM_C030/2020|EPI_ISL_434376|2020- ···················· ···················· ···················· ···················· NNNNNNNNNNNNNNNNNNNN NNNNNNNNNNNNNNNNNNNN 1
B 6998 hCoV-19/Belgium/ITM_C451/2020|EPI_ISL_434377|2020- ···················· ···················· ···················· ···················· NNNNNNNNNNNNNNNNNNNN NNNNNNNNNNNNNNNNNNNN 1
B 6999 hCoV-19/Belgium/ITM_C456/2020|EPI_ISL_434378|2020- ···················· ···················· ···················· ···················· NNNNNNNNNNNNNNNNNNNN NNNNNNNNNNNNNNNNNNNN 1
B 7000 hCoV-19/Belgium/ITM_C285/2020|EPI_ISL_434379|2020- ···················· ···················· ···················· ···················· NNNNNNNNNNNNNNNNNNNN NNNNNNNNNNNNNNNNNNNN 1
B 7013 hCoV-19/USA/CA-SCCPHD-UC178/2020|EPI_ISL_435657|20 ···················· ···················· ···················· ···················· ···················· ···NNNNNNNNNNNNNNNNN 1
B 7788 hCoV-19/England/CAMB-7857D/2020|EPI_ISL_433749|202 ···················· ···················· ···················· ···················· NNNNNNNNNNNNNNNNNNNN NNNNNNNNNNNNNNNNNNNN 1
B 7902 hCoV-19/England/BIRM-5E2FE/2020|EPI_ISL_444284|202 ···················· ···················· ···················· ···················· NNNNNNNNNNNNNNNNNNNN NNNNNNNNNNNNNNNNNNNN 1
B 7907 hCoV-19/England/BIRM-5E373/2020|EPI_ISL_444290|202 ···················· ···················· ···················· ···················· NNNNNNNNNNNNNNNNNNNN NNNNNNNNNNNNNNNNNNNN 1
B 7911 hCoV-19/England/BIRM-5E6B6/2020|EPI_ISL_444299|202 ···················· ···················· ···················· ···················· NNNNNNNNNNNNNNNNNNNN NNNNNNNNNNNNNNNNNNNN 1
B 7962 hCoV-19/England/CAMB-78328/2020|EPI_ISL_433718|202 ···················· ···················· ···················· ···················· NNNNNNNNNNNNNNNNNNNN NNNNNNNNNNNNNNNNNNNN 1
B 7963 hCoV-19/England/CAMB-78337/2020|EPI_ISL_433719|202 ···················· ···················· ···················· ···················· NNNNNNNNNNNNNNNNNNNN NNNNNNNNNNNNNNNNNNNN 1
B 8036 hCoV-19/Germany/BY-MVP-0069/2020|EPI_ISL_437265|20 ···················· ···················· ···················· ···················· ····NNNNNNNNNNNNNNNN N···NNNNNNNNNNN····· 1
B 8056 hCoV-19/Germany/BY-MVP-0091/2020|EPI_ISL_437285|20 ···················· ···················· ···················· ···················· ····NNNNNNNNNNNNNNNN ···················· 1
```

| | | | | | | | | | |
|---|---|---|---|---|---|---|---|---|---|
| B | 8064 | hCoV-19/Germany/BY-MVP-0104/2020\|EPI_ISL_437294\|20 | ···················· | ···················· | ···················· | ···················· | NNNNNNNNNNNNNNNNNNNN | ···················· | 1 |
| B | 8065 | hCoV-19/Germany/BY-MVP-0105/2020\|EPI_ISL_437295\|20 | ···················· | ···················· | ···················· | ···················· | NNNNNNNNNNNNNNNNNNNN | ···················· | 1 |
| B | 8192 | hCoV-19/Australia/VIC770/2020\|EPI_ISL_450214\|2020- | ···················· | ···················· | ···················· | ···················· | NNNNNNNNNNNNNNNNNNNN | NNNNNNNNNNNNNNNNNNNN | 1 |
| B | 8258 | hCoV-19/Germany/BY-MVP-0036/2020\|EPI_ISL_437235\|20 | ···················· | ···················· | ···················· | ···················· | ···············NNNNN | ···················· | 1 |
| B | 8264 | hCoV-19/Germany/BY-MVP-0043/2020\|EPI_ISL_437242\|20 | ···················· | ···················· | ···················· | ···················· | NNNNNNNNNNNNNNNNNNNN | NNNNNNNNNNNNNNNNN··· | 1 |
| B | 8498 | hCoV-19/Myanmar/NIH-4385/2020\|EPI_ISL_434709\|2020 | ···················· | ···················· | ···················· | ···················· | ······NNNNNNNNNNNNNN | ···················· | 1 |
| B | 8519 | hCoV-19/USA/TX-HMH026/2020\|EPI_ISL_434735\|2020-03- | ···················· | ···················· | ···················· | ···················· | NNNNNNNNNNNNNNNNNNNN | NNNNNNNNNNNNNNNNNNNN | 1 |
| B | 8523 | hCoV-19/USA/TX-HMH037/2020\|EPI_ISL_434746\|2020-03- | ···················· | ···················· | ···················· | ···················· | NNNNNNNNNNNNNNNNNNNN | NNNNNNNNNNNNNNNNNNNN | 1 |
| B | 8525 | hCoV-19/USA/TX-HMH046/2020\|EPI_ISL_434755\|2020-03- | ···················· | ···················· | ···················· | ···················· | NNNNNNNNNNNNNNNNNNNN | NNNNNNNNNNNNNNNNNNNN | 1 |
| B | 8527 | hCoV-19/USA/TX-HMH051/2020\|EPI_ISL_434760\|2020-03- | ···················· | N··················· | ···················· | ···················· | NNNNNNNNNNNNNNNNNNNN | NNNNNNNNNNNNNNNNNNNN | 1 |
| B | 8529 | hCoV-19/USA/TX-HMH057/2020\|EPI_ISL_434766\|2020-03- | ···················· | ···················· | ···················· | ···················· | NNNNNNNNNNNNNNNNNNNN | NNNNNNNNNNNNNNNNNNNN | 1 |
| B | 8530 | hCoV-19/USA/TX-HMH059/2020\|EPI_ISL_434768\|2020-03- | ···················· | ···················· | ···················· | ···················· | NNNNNNNNNNNNNNNNNNNN | NNNNNNNNNNNNNNNNNNNN | 1 |
| B | 8547 | hCoV-19/USA/MD-HP00087/2020\|EPI_ISL_438241\|2020-03 | ···················· | ···················· | ···················· | ···················· | NNNNNNNNNNNNNNNNNNNN | NNNNNNNNNNNNNNNNNNNN | 1 |
| B | 8548 | hCoV-19/USA/DC-HP00089/2020\|EPI_ISL_438242\|2020-03 | ···················· | ···················· | ···················· | ···················· | NNNNNNNNNNNNNNNNNNNN | NNNNNNNNNNNNNNNNNNNN | 1 |
| B | 8549 | hCoV-19/USA/DC-HP00090/2020\|EPI_ISL_438243\|2020-03 | ···················· | ···················· | ···················· | ···················· | NNNNNNNNNNNNNNNNNNNN | NNNNNNNNNNNNNNNNNNNN | 1 |
| B | 8550 | hCoV-19/USA/DC-HP00099/2020\|EPI_ISL_438244\|2020-03 | ···················· | ···················· | ···················· | ···················· | NNNNNNNNNNNNNNNNNNNN | NNNNNNNNNNNNNNNNNNNN | 1 |
| B | 8551 | hCoV-19/USA/DC-HP00100/2020\|EPI_ISL_438245\|2020-03 | ···················· | ···················· | ···················· | ···················· | NNNNNNNNNNNNNNNNNNNN | NNNNNNNNNNNNNNNNNNNN | 1 |
| B | 8553 | hCoV-19/USA/MD-HP00105/2020\|EPI_ISL_438247\|2020-03 | ···················· | ···················· | ···················· | ···················· | NNNNNNNNNNNNNNNNNNNN | NNNNNNNNNNNNNNNNNNNN | 1 |
| B | 9073 | hCoV-19/USA/TX-HMH0105/2020\|EPI_ISL_434810\|2020-03 | ···················· | ···················· | ···················· | ···················· | NNNNNNNNNNNNNNNNNNNN | NNNNNNNNNNNNNNNNNNNN | 1 |
| B | 9076 | hCoV-19/USA/TX-HMH0111/2020\|EPI_ISL_434813\|2020-03 | ···················· | ···················· | ···················· | ···················· | NNNNNNNNNNNNNNNNNNNN | NNNNNNNNNNNNNNNNNNNN | 1 |
| B | 9077 | hCoV-19/USA/TX-HMH0113/2020\|EPI_ISL_434815\|2020-03 | ···················· | ···················· | ···················· | ···················· | NNNNNNNNNNNNNNNNNNNN | NNNNNNNNNNNNNNNNNNNN | 1 |
| B | 9087 | hCoV-19/USA/TX-HMH0133/2020\|EPI_ISL_434833\|2020-03 | ···················· | ···················· | ···················· | ···················· | N·············N····· | NN·N·NNNNNNN········ | 1 |
| B | 9089 | hCoV-19/USA/TX-HMH0137/2020\|EPI_ISL_434837\|2020-03 | ···················· | ···················· | ···················· | ···················· | ·N·N··········NN·N·· | N······NN·····N····· | 1 |
| B | 9211 | hCoV-19/Wales/PHWC-32E74/2020\|EPI_ISL_446699\|2020- | ···················· | ·A·················· | ···················· | ···················· | NNNN················ | ···················· | 1 |
| B | 9256 | hCoV-19/USA/TX-HMH097/2020\|EPI_ISL_434802\|2020-03- | ···················· | ···················· | ···················· | ···················· | NNNNNNNNNNNNNNNNNNNN | NNNNNNNNNNNNNNNNNNNN | 1 |
| B | 9260 | hCoV-19/USA/TX-HMH0103/2020\|EPI_ISL_434808\|2020-03 | ···················· | ···················· | ···················· | ···················· | ···················· | N··N···NN··········· | 1 |
| B | 9512 | hCoV-19/USA/TX-HMH068/2020\|EPI_ISL_434777\|2020-03- | ···················· | ···················· | ···················· | ···················· | NNNNNNNNNNNNNNNNNNNN | NNNNNNNNNNNNNNNNNNNN | 1 |
| B | 9514 | hCoV-19/USA/TX-HMH072/2020\|EPI_ISL_434781\|2020-03- | ···················· | ···················· | ···················· | ···················· | NNNNNNNNNNNNNNNNNNNN | NNNNNNNNNNNNNNNNNNNN | 1 |
| B | 9516 | hCoV-19/USA/TX-HMH075/2020\|EPI_ISL_434784\|2020-03- | ···················· | ···················· | ···················· | ···················· | NNNNNNNNNNNNNNNNNNNN | NNNNNNNNNNNNNNNNNNNN | 1 |
| B | 9520 | hCoV-19/USA/TX-HMH081/2020\|EPI_ISL_434790\|2020-03- | ···················· | ···················· | ···················· | ···················· | NNNNNNNNNNNNNNNNNNNN | NNNNNNNNNNNNNNNNNNNN | 1 |
| B | 9521 | hCoV-19/USA/TX-HMH082/2020\|EPI_ISL_434791\|2020-03- | ···················· | ···················· | ···················· | ···················· | NNNNNNNNNNNNNNNNNNNN | NNNNNNNNNNNNNNNNNNNN | 1 |
| B | 9522 | hCoV-19/USA/TX-HMH084/2020\|EPI_ISL_434792\|2020-03- | ···················· | ···················· | ···················· | ···················· | NNNNNNNNNNNNNNNNNNNN | NNNNNNNNNNNNNNNNNNNN | 1 |
| B | 9523 | hCoV-19/USA/TX-HMH089/2020\|EPI_ISL_434795\|2020-03- | ···················· | ···················· | ···················· | ···················· | NNNNNNNNNNNNNNNNNNNN | NNNNNNNNNNNNNNNNNNNN | 1 |
| B | 9524 | hCoV-19/USA/TX-HMH092/2020\|EPI_ISL_434797\|2020-03- | ···················· | ···················· | ···················· | ···················· | ·············NNN···· | NN·····NN·NN···N···· | 1 |
| B | 9526 | hCoV-19/USA/TX-HMH094/2020\|EPI_ISL_434799\|2020-03- | ···················· | ···················· | ···················· | ···················· | NNNNNNNNNNNNNNNNNNNN | NNNNNNNNNNNNNNNNNNNN | 1 |
| B | 9604 | hCoV-19/Scotland/EDB3944/2020\|EPI_ISL_439669\|2020- | ···················· | ·A·················· | ···················· | ···················· | NNNNNNNNNNNNNNNNNNNN | NNNNNNNNNNNNNNNNNNNN | 1 |
| C | 57 | hCoV-19/Switzerland/BL-ETHZ-110430/2020\|EPI_ISL_45 | ···················· | ···················· | ···················· | ···················· | ······NNNNNNNNNNNNNN | NNNNNNNNNNNNNNNNNNNN | 1 |
| C | 81 | hCoV-19/Nigeria/OS016-CV3/2020\|EPI_ISL_455413\|2020 | ·NNNNNNNNNNNNNNNNNNN | NNNNNNNNNNNNNNNNNNNN | ···················· | ···················· | ···················· | ···················· | 1 |
| C | 92 | hCoV-19/Mexico/CMX-InDRE_12/2020\|EPI_ISL_455436\|20 | ···················· | ···················· | ···················· | ···················· | NNNNNNNNNNN········· | ···················· | 1 |
| C | 155 | hCoV-19/England/CAMB-1B31DB/2020\|EPI_ISL_456751\|20 | ···················· | ···················· | ···················· | ···················· | NNNNNNNNNNNNNNNNNNNN | NNNNNNNNNNNNNNNNNNNN | 1 |
| C | 158 | hCoV-19/Scotland/CVR1051/2020\|EPI_ISL_456756\|2020- | ···················· | ···················· | ···················· | ···················· | NNNNNNNNNNNNNNNNNNNN | ···················· | 1 |
| C | 159 | hCoV-19/Scotland/CVR1891/2020\|EPI_ISL_456757\|2020- | ···················· | ···················· | ···················· | ···················· | NNNNNNNNNNNNNNNNNNNN | ···················· | 1 |
| C | 166 | hCoV-19/Scotland/CVR3277/2020\|EPI_ISL_456774\|2020- | ···················· | ···················· | ···················· | ···················· | NNNNNNNNNNNNNNNNNNNN | ···················· | 1 |
| C | 170 | hCoV-19/Scotland/CVR3298/2020\|EPI_ISL_456779\|2020- | ···················· | ···················· | ···················· | ···················· | NNNNNNNNNNNNNNNNNNNN | NNNNNNNNNNNNNNNNNNNN | 1 |
| C | 174 | hCoV-19/Scotland/CVR3347/2020\|EPI_ISL_456783\|2020- | ···················· | ···················· | ···················· | ···················· | NNNNNNNNNNNNNNNNNNNN | ···················· | 1 |
| C | 177 | hCoV-19/Scotland/CVR3354/2020\|EPI_ISL_456786\|2020- | ···················· | ···················A | ···················· | ···················· | NNNNNNNNNNNNNNNNNNNN | ···················· | 1 |
| C | 178 | hCoV-19/Scotland/CVR3355/2020\|EPI_ISL_456787\|2020- | ···················· | ···················· | ···················· | ···················· | NNNNNNNNNNNNNNNNNNNN | ···················· | 1 |
| C | 185 | hCoV-19/Scotland/CVR3364/2020\|EPI_ISL_456794\|2020- | ···················· | ···················· | ···················· | ···················· | NNNNNNNNNNNNNNNNNNNN | ···················· | 1 |
| C | 187 | hCoV-19/Scotland/CVR3366/2020\|EPI_ISL_456796\|2020- | ···················· | ···················A | ···················· | ···················· | NNNNNNNNNNNNNNNNNNNN | ···················· | 1 |
| C | 188 | hCoV-19/Scotland/CVR3367/2020\|EPI_ISL_456797\|2020- | ···················· | ···················· | ···················· | ···················· | ······NNNNNNNNNNNNNN | ···················· | 1 |
| C | 275 | hCoV-19/England/NORW-E8709/2020\|EPI_ISL_448226\|202 | ···················· | ···················· | ···················· | ···················· | ···················· | NNNN···NN··········· | 1 |
| C | 304 | hCoV-19/England/NORW-E89B8/2020\|EPI_ISL_448258\|202 | ···················· | ···················· | ···················· | ···················· | NNNNNNNNNNNNN······· | NNNNNNNNN··········· | 1 |
| C | 340 | hCoV-19/Switzerland/SG-ETHZ-101137/2020\|EPI_ISL_45 | ···················· | ···················· | ···················· | ···················· | ······NNNNNNNNNNNNNN | NNNNNNNNNNNNNNNNNNNN | 1 |
| C | 345 | hCoV-19/Switzerland/BS-ETHZ-101147/2020\|EPI_ISL_45 | ···················· | ···················· | ···················· | ···················· | ······NNNNNNNNNNNNNN | NNNNNNNNNNNNNNNNNNNN | 1 |
| C | 346 | hCoV-19/Switzerland/BL-ETHZ-101148/2020\|EPI_ISL_45 | ···················· | ···················· | ···················· | ···················· | ······NNNNNNNNNNNNNN | NNNNNNNNNNNNNNNNNNNN | 1 |
| C | 347 | hCoV-19/Switzerland/BL-ETHZ-101149/2020\|EPI_ISL_45 | ···················· | ···················· | ···················· | ···················· | ······SNNNNNNNNNNNNN | ···················· | 1 |
| C | 349 | hCoV-19/Switzerland/VS-ETHZ-101151/2020\|EPI_ISL_45 | ···················· | ···················· | ···················· | ···················· | ······NNNNNNNNNNNNNN | NNNNNNNNNNNNNNNNNNNN | 1 |
| C | 353 | hCoV-19/Switzerland/AG-ETHZ-101157/2020\|EPI_ISL_45 | ···················· | ···················A | ···················· | ···················· | ······NNNNNNNNNNNNNN | ···················· | 1 |
| C | 356 | hCoV-19/Switzerland/LU-ETHZ-101161/2020\|EPI_ISL_45 | ···················· | ···················· | ···················· | ···················· | ······NNNNNNNNNNNNNN | ···················· | 1 |
| C | 358 | hCoV-19/Switzerland/BE-ETHZ-101166/2020\|EPI_ISL_45 | ···················· | ···················· | ···················· | ···················· | ······NNNNNNNNNNNNNN | NNNNNNNNNNNNNNNNNNNN | 1 |
| C | 359 | hCoV-19/Switzerland/ZH-ETHZ-101167/2020\|EPI_ISL_45 | ···················· | ···················· | ···················· | ···················· | ······NNNNNNNNNNNNNN | ···················· | 1 |
| C | 365 | hCoV-19/Switzerland/ZH-ETHZ-101175/2020\|EPI_ISL_45 | ···················· | ···················· | ···················· | ···················· | ······NNNNNNNNNNNNNN | ···················· | 1 |
| C | 368 | hCoV-19/Switzerland/ZH-ETHZ-101180/2020\|EPI_ISL_45 | ···················· | ···················· | ···················· | ···················· | ······NNNNNNNNNNNNNN | ···················· | 1 |
| C | 373 | hCoV-19/Switzerland/JU-ETHZ-101187/2020\|EPI_ISL_45 | ···················· | ···················· | ···················· | ···················· | ······NNNNNNNNNNNNNN | ···················· | 1 |
| C | 374 | hCoV-19/Switzerland/GR-ETHZ-101188/2020\|EPI_ISL_45 | ···················· | ···················· | ···················· | ···················· | ······NNNNNNNNNNNNNN | ···················· | 1 |
| C | 375 | hCoV-19/Switzerland/GE-ETHZ-101189/2020\|EPI_ISL_45 | ···················· | ···················· | ···················· | ···················· | ······NNNNNNNNNNNNNN | NNNNNNNNNNNNNNNNNNNN | 1 |
| C | 380 | hCoV-19/Switzerland/ZH-ETHZ-110436/2020\|EPI_ISL_45 | ···················· | ···················· | ···················· | ···················· | ······NNNNNNNNNNNNNN | NNNNNNNNNNNNNNNNNNNN | 1 |
| C | 381 | hCoV-19/Switzerland/ZH-ETHZ-110437/2020\|EPI_ISL_45 | ···················· | ···················· | ···················· | ···················· | ······NNNNNNNNNNNNNN | NNNNNNNNNNNNNNNNNNNN | 1 |
| C | 382 | hCoV-19/Switzerland/ZH-ETHZ-110439/2020\|EPI_ISL_45 | ···················· | ···················· | ···················· | ···················· | ······NNNNNNNNNNNNNN | NNNNNNNNNNNNNNNNNNNN | 1 |
| C | 383 | hCoV-19/Switzerland/AG-ETHZ-110444/2020\|EPI_ISL_45 | ···················· | ···················· | ···················· | ···················· | ······NNNNNNNNNNNNNN | NNNNNNNNNNNNNNNNNNNN | 1 |
| C | 384 | hCoV-19/Switzerland/VD-ETHZ-110448/2020\|EPI_ISL_45 | ···················· | ···················· | ···················· | ···················· | ······NNNNNNNNNNNNNN | NNNNNNNNNNNNNNNNNNNN | 1 |
| C | 385 | hCoV-19/Switzerland/GR-ETHZ-110450/2020\|EPI_ISL_45 | ···················· | ···················· | ···················· | ···················· | ······NNNNNNNNNNNNNN | ···················· | 1 |
| C | 386 | hCoV-19/Switzerland/LU-ETHZ-110451/2020\|EPI_ISL_45 | ···················· | ···················· | ···················· | ···················· | ······NNNNNNNNNNNNNN | NNNNNNNNNNNNNNNNNNNN | 1 |
| C | 387 | hCoV-19/Switzerland/LU-ETHZ-110452/2020\|EPI_ISL_45 | ···················· | ···················· | ···················· | ···················· | ······NNNNNNNNNNNNNN | ···················· | 1 |
| C | 388 | hCoV-19/Switzerland/LU-ETHZ-110453/2020\|EPI_ISL_45 | ···················· | ···················· | ···················· | ···················· | ······NNNNNNNNNNNNNN | NNNNNNNNNNNNNNNNNNNN | 1 |
| C | 389 | hCoV-19/Switzerland/BS-ETHZ-110454/2020\|EPI_ISL_45 | ···················· | ·A·················· | ···················· | ···················· | ······NNNNNNNNNNNNNN | NNNNNNNNNNNNNNNNNNNN | 1 |
| C | 391 | hCoV-19/Switzerland/BE-ETHZ-110458/2020\|EPI_ISL_45 | ···················· | ···················· | ···················· | ···················· | ······NNNNNNNNNNNNNN | NNNNNNNNNNNNNNNNNNNN | 1 |
| C | 392 | hCoV-19/Switzerland/GL-ETHZ-110459/2020\|EPI_ISL_45 | ···················· | ···················· | ···················· | ···················· | ······NNNNNNNNNNNNNN | NNNNNNNNNNNNNNNNNNNN | 1 |
| C | 393 | hCoV-19/Switzerland/BE-ETHZ-110460/2020\|EPI_ISL_45 | ···················· | ···················· | ···················· | ···················· | ······NNNNNNNNNNNNNN | NNNNNNNNNNNNNNNNNNNN | 1 |
| C | 394 | hCoV-19/Switzerland/BL-ETHZ-110465/2020\|EPI_ISL_45 | ···················· | ···················· | ···················· | ···················· | ······NNNNNNNNNNNNNN | NNNNNNNNNNNNNNNNNNNN | 1 |

```
C   395 hCoV-19/Switzerland/VD-ETHZ-110466/2020|EPI_ISL_45   ···················· ···················· ···················· ···················· ·······NNNNNNNNNNNNN NNNNNNNNNNNNNNNNNNNN   1
C   396 hCoV-19/Switzerland/BS-ETHZ-110467/2020|EPI_ISL_45   ···················· ···················· ···················· ···················· ·······NNNNNNNNNNNNN NNNNNNNNNNNNNNNNNNNN   1
C   397 hCoV-19/Switzerland/BE-ETHZ-110468/2020|EPI_ISL_45   ···················· ···················· ···················· ···················· ·······NNNNNNNNNNNNN NNNNNNNNNNNNNNNNNNNN   1
C   398 hCoV-19/Switzerland/VD-ETHZ-110469/2020|EPI_ISL_45   ···················· ·······Y············ ···················· ···················· ·······NNNNNNNNNNNNN NNNNNNNNNNNNNNNNNNNN   1
C   400 hCoV-19/Switzerland/FR-ETHZ-110474/2020|EPI_ISL_45   ···················· ···················· ···················· ···················· ·······NNNNNNNNNNNNN ····················   1
C   401 hCoV-19/Switzerland/TI-ETHZ-110479/2020|EPI_ISL_45   ···················· ···················· ···················· ···················· ·······NNNNNNNNNNNNN ····················   1
C   402 hCoV-19/Switzerland/BL-ETHZ-110480/2020|EPI_ISL_45   ···················· ···················· ···················· ···················· ·······NNNNNNNNNNNNN ····················   1
C   403 hCoV-19/Switzerland/BL-ETHZ-110482/2020|EPI_ISL_45   ···················· ···················· ···················· ···················· ·······NNNNNNNNNNNNN ····················   1
C   492 hCoV-19/England/CAMB-1B250D/2020|EPI_ISL_456688|20   ···················· ···················· ···················· ···················· NNNNNNNNNNNNNNNNNNNN NNNNNNNNNNNNNNNNNNNN   1
C   493 hCoV-19/England/CAMB-1B25C1/2020|EPI_ISL_456697|20   ···················· ···················· ···················· ···················· NNNNNNNNNNNNNNNNNNNN NNNNNNNNNNNNNNNNNNNN   1
C   527 hCoV-19/Switzerland/ZH-ETHZ-101134/2020|EPI_ISL_45   ···················· ···················· ···················· ···················· ·······NNNNNNNNNNNNN NNNNNNNNNNNNNNNNNNNN   1
C   531 hCoV-19/Switzerland/LU-ETHZ-101141/2020|EPI_ISL_45   ···················· ···················· ···················· ···················· ·······SNNNNNNNNNNNN ····················   1
C   640 hCoV-19/Scotland/CVR3409/2020|EPI_ISL_456833|2020-   ···················· ···················A ···················· ···················· NNNNNNNNNNNNNNNNNNNN ····················   1
C   643 hCoV-19/Scotland/CVR3415/2020|EPI_ISL_456838|2020-   ···················· ···················· ···················· ···················· NNNNNNNNNNNNNNNNNNNN ····················   1
C   645 hCoV-19/Scotland/CVR3417/2020|EPI_ISL_456840|2020-   ···················· ··············C····· ···················· ···················· NNNNNNNNNNNNNNNNNNNN NNNNNNNNNNNNNNNNNNNN   1
C   646 hCoV-19/Scotland/CVR3421/2020|EPI_ISL_456842|2020-   ···················· ···················A ···················· ···················· NNNNNNNNNNNNNNNNNNNN ····················   1
C   647 hCoV-19/Scotland/CVR3426/2020|EPI_ISL_456847|2020-   ···················· ···················A ···················· ···················· NNNNNNNNNNNNNNNNNNNN ····················   1
C   648 hCoV-19/Scotland/CVR3427/2020|EPI_ISL_456848|2020-   ···················· ···················A ···················· ···················· ········NNNNNNNNNNNN ····················   1
C   649 hCoV-19/Scotland/CVR3436/2020|EPI_ISL_456851|2020-   ···················· ···················· ···················· ···················· NNNNNNNNNNNNNNNNNNNN ····················   1
C   650 hCoV-19/Scotland/CVR3439/2020|EPI_ISL_456852|2020-   ···················· ···················· ···················· ···················· NNNNNNNNNNNNNNNNNNNN ····················   1
C   651 hCoV-19/Scotland/CVR3441/2020|EPI_ISL_456854|2020-   ···················· ···················· ···················· ···················· NNNNNNNNNNNNNNNNNNNN NN··················   1
C   653 hCoV-19/Scotland/CVR3448/2020|EPI_ISL_456856|2020-   ···················· ···················· ···················· ···················· NNNNNNNNNNNNNNNNNNNN ····················   1
C   659 hCoV-19/Scotland/CVR3477/2020|EPI_ISL_456870|2020-   ···················· ···················A ···················· ···················· NNNNNNNNNNNNNNNNNNNN ····················   1
C   660 hCoV-19/Scotland/CVR3478/2020|EPI_ISL_456871|2020-   ···················· ···················A ···················· ···················· NNNNNNNNNNNNNNNNNNNN NNNNNNNNNNNNNNNNNNNN   1
C   661 hCoV-19/Scotland/CVR3479/2020|EPI_ISL_456872|2020-   ···················· ···················A ···················· ···················· NNNNNNNNNNNNNNNNNNNN ····················   1
C   668 hCoV-19/Scotland/CVR3499/2020|EPI_ISL_456884|2020-   ···················· ···················· ···················· ···················· NNNNNNNNNNNNNNNNNNNN ····················   1
C   669 hCoV-19/Scotland/CVR3500/2020|EPI_ISL_456885|2020-   ···················· ···················· ···················· ···················· NNNNNNNNNNNNNNNNNNNN NNNNNNNNNNNNNNNNNNNN   1
C   776 hCoV-19/England/NORW-E93C8/2020|EPI_ISL_448309|202   ···················· ···················· ···················· ···················· NNNNNNNNNNNNNNNNNNNN NNNNNNNNNNNNNNNNNNNN   1
C   787 hCoV-19/England/NORW-E961A/2020|EPI_ISL_448323|202   ···················· ···················· ···················· ···················· NNNNNNNNNNNNNNNNNNNN NNNNNNNNNNNNNNNNNNNN   1
C   789 hCoV-19/England/NORW-E9683/2020|EPI_ISL_448325|202   ···················· ···················· ···················· ···················· ·NN····NNNNN·······N ····················   1
C   794 hCoV-19/England/NORW-E9762/2020|EPI_ISL_448330|202   ···················· ···················· ···················· ···················· NNNNNNNNNNNNNNNNNNNN ····················   1
C   816 hCoV-19/England/NORW-E9ED6/2020|EPI_ISL_448379|202   ···················· ···················· ···················· ···················· NNNNNNNNNNNNNNNNNNNN NNNN················   1
C   902 hCoV-19/Scotland/CVR3382/2020|EPI_ISL_456811|2020-   ···················· ·A·················· ···················· ···················· NNNNNNNNNNNNNNNNNNNN ····················   1
C   904 hCoV-19/Scotland/CVR3387/2020|EPI_ISL_456815|2020-   ···················· ···················· ···················· ···················· NNNNNNNNNNNNNNNNNNNN ····················   1
C   905 hCoV-19/Scotland/CVR3390/2020|EPI_ISL_456818|2020-   ···················· ···················· ···················· ···················· NNNNNNNNNNNNNNNNNNNN ····················   1
C   907 hCoV-19/Scotland/CVR3394/2020|EPI_ISL_456821|2020-   ···················· ··············C····· ···················· ···················· NNNNNNNNNNNNNNNNNNNN ····················   1
C   908 hCoV-19/Scotland/CVR3397/2020|EPI_ISL_456824|2020-   ···················· ···················· ···················· ···················· NNNNNNNNNNNNNNNNNNNN NNNNNNNNNNNNNNNNNNNN   1
C   909 hCoV-19/Scotland/CVR3400/2020|EPI_ISL_456826|2020-   ···················· ···················A ···················· ···················· NNNNNNNNNNNNNNNNNNNN ····················   1
C   913 hCoV-19/Scotland/CVR3407/2020|EPI_ISL_456831|2020-   ···················· ···················A ···················· ···················· NNNNNNNNNNNNNNNNNNNN NNNNNNNNNNNNNNNNNNNN   1
C  1065 hCoV-19/Switzerland/BL-ETHZ-110489/2020|EPI_ISL_45   ···················· ··············Y····· ···················· ···················· ·······NNNNNNNNNNNNN NNNNNNNNNNNNNNNNNNNN   1
C  1066 hCoV-19/Switzerland/BE-ETHZ-110491/2020|EPI_ISL_45   ···················· ···················· ···················· ···················· ·······NNNNNNNNNNNNN NNNNNNNNNNNNNNNNNNNN   1
C  1067 hCoV-19/Switzerland/BE-ETHZ-110497/2020|EPI_ISL_45   ···················· ··········M········· ···················· ···················· ·······NNNNNNNNNNNNN NNNNNNNNNNNNNNNNNNNN   1
C  1068 hCoV-19/Switzerland/ZH-ETHZ-110498/2020|EPI_ISL_45   ···················· ···················· ···················· ···················· ·······NNNNNNNNNNNNN NNNNNNNNNNNNNNNNNNNN   1
C  1069 hCoV-19/Switzerland/BE-ETHZ-110499/2020|EPI_ISL_45   ···················· ·········K·········· ···················· ···················· ·······NNNNNNNNNNNNN ····················   1
C  1070 hCoV-19/Switzerland/BE-ETHZ-110500/2020|EPI_ISL_45   ···················· ···················· ···················· ···················· ·······NNNNNNNNNNNNN ····················   1
C  1071 hCoV-19/Switzerland/AG-ETHZ-110503/2020|EPI_ISL_45   ···················· ···················· ···················· ···················· ·······NNNNNNNNNNNNN NNNNNNNNNNNNNNNNNNNN   1
C  1072 hCoV-19/Switzerland/ZH-ETHZ-110505/2020|EPI_ISL_45   ···················· ···················· ···················· ···················· ·······NNNNNNNNNNNNN ····················   1
C  1073 hCoV-19/Switzerland/VD-ETHZ-110507/2020|EPI_ISL_45   ···················· ···············R···· ···················· ···················· ·······NNNNNNNNNNNNN NNNNNNNNNNNNNNNNNNNN   1
C  1075 hCoV-19/Switzerland/BL-ETHZ-110509/2020|EPI_ISL_45   ···················· ···················· ···················· ···················· ·······NNNNNNNNNNNNN NNNNNNNNNNNNNNNNNNNN   1
C  1076 hCoV-19/Switzerland/AG-ETHZ-110510/2020|EPI_ISL_45   ···················· ···················· ···················· ···················· ·······NNNNNNNNNNNNN NNNNNNNNNNNNNNNNNNNN   1
C  1168 hCoV-19/Australia/VIC1581/2020|EPI_ISL_456500|2020   ···················· ···················· ···················· ···················· NNNNNNNNNNNNNNNNNNNN NNNNNNNNNNNNNNNNNNNN   1
C  1169 hCoV-19/Australia/VIC1582/2020|EPI_ISL_456501|2020   ···················· ···················· ···················· ···················· NNNNNNNNNNNNNNNNNNNN NNNNNNNNNNNNNNNNNNNN   1
C  1170 hCoV-19/Australia/VIC1583/2020|EPI_ISL_456502|2020   ···················· ···················· ···················· ···················· NNNNNNNNNNNNNNNNNNNN NNNNNNNNNNNNNNNNNNNN   1
C  1171 hCoV-19/Australia/VIC1584/2020|EPI_ISL_456503|2020   ···················· ···················· ···················· ···················· NNNNNNNNNNNNNNNNNNNN NNNNNNNNNNNNNNNNNNNN   1
C  1172 hCoV-19/Australia/VIC1585/2020|EPI_ISL_456504|2020   ···················· ···················· ···················· ···················· ·······NNNNNNNNNNNNN NNNNNNNNNNNNNNNNNNNN   1
C  1173 hCoV-19/Australia/VIC1586/2020|EPI_ISL_456506|2020   ···················· ···················· ···················· ···················· NNNNNNNNNNNNNNNNNNNN NNNNNNNNNNNNNNNNNNNN   1
C  1174 hCoV-19/Australia/VIC1587/2020|EPI_ISL_456507|2020   ···················· ···················· ···················· ···················· NNNNNNNNNNNNNNNNNNNN NNNNNNNNNNNNNNNNNNNN   1
C  1175 hCoV-19/Australia/VIC1588/2020|EPI_ISL_456508|2020   ···················· ···················· ···················· ···················· NNNNNNNNNNNNNNNNNNNN NNNN················   1
C  1176 hCoV-19/Australia/VIC1589/2020|EPI_ISL_456509|2020   ···················· ···················· ···················· ···················· NNNNNNNNNNNNNNNNNNNN NNNNNNNNNNNNNNNNNNNN   1
C  1177 hCoV-19/Australia/VIC1590/2020|EPI_ISL_456510|2020   ···················· ···················· ···················· ···················· NNNNNNNNNNNNNNNNNNNN NNNNNNNNNNNNNNNNNNNN   1
C  1178 hCoV-19/Australia/VIC1591/2020|EPI_ISL_456511|2020   ···················· ···················· ···················· ···················· NNNNNNNNNNNNNNNNNNNN NNNNNNNNNNNNNNNNNNNN   1
C  1179 hCoV-19/Australia/VIC1592/2020|EPI_ISL_456512|2020   ···················· ···················· ···················· ···················· NNNNNNNNNNNNNNNNNNNN NNNNNNNNNNNNNNNNNNNN   1
C  1180 hCoV-19/Australia/VIC1631/2020|EPI_ISL_456536|2020   ···················· ···················· ···················· ···················· NNNNNNNNNNNNNNNNNNNN NNNNNNNNNNNNNNNNNNNN   1
C  1181 hCoV-19/Australia/VIC1633/2020|EPI_ISL_456538|2020   ···················· ···················· ···················· ···················· NNNNNNNNNNNNNNNNNNNN NNNNNNNNNNNNNNNNNNNN   1
C  1182 hCoV-19/Australia/VIC1634/2020|EPI_ISL_456539|2020   ···················· ···················· ···················· ···················· NNNNNNNNNNNNNNNNNNNN NNNNNNNNNNNNNNNNNNNN   1
C  1183 hCoV-19/Australia/VIC1637/2020|EPI_ISL_456540|2020   ···················· ···················· ···················· ···················· NNNNNNNNNNNNNNNNNNNN NNNNNNNNNNNNNNNNNNNN   1
C  1184 hCoV-19/Australia/VIC1638/2020|EPI_ISL_456541|2020   ···················· ···················· ···················· ···················· NNNNNNNNNNNNNNNNNNNN NNNNNNNNNNNNNNNNNNNN   1
C  1185 hCoV-19/Australia/VIC1646/2020|EPI_ISL_456548|2020   ···················· ···················· ···················· ···················· NNNNNNNNNNNNNNNNNNNN NNNNNNNNNNNNNNNNNNNN   1
C  1186 hCoV-19/Australia/VIC1648/2020|EPI_ISL_456550|2020   ···················· ···················· ···················· ···················· NNNNNNNNNNNNNNNNNNNN NNNNNNNNNNNNNNNNNNNN   1
C  1187 hCoV-19/Australia/VIC1651/2020|EPI_ISL_456553|2020   ···················· ···················· ···················· ···················· NNNNNNNNNNNNNNNNNNNN NNNNNNNNNNNNNNNNNNNN   1
C  1188 hCoV-19/Australia/VIC1653/2020|EPI_ISL_456555|2020   ···················· ···················· ···················· ···················· NNNNNNNNNNNNNNNNNNNN NNNNNNNNNNNNNNNNNNNN   1
C  1192 hCoV-19/USA/MD-HP00060/2020|EPI_ISL_457802|2020-03   ···················· ···················· ···················· ···················· NNNNNNNNNNNNNNNNNNNN NNNNNNNNNNNNNNNNNNNN   1
C  1194 hCoV-19/USA/MD-HP00062/2020|EPI_ISL_457804|2020-03   ···················· ···················· ···················· ···················· NNNNNNNNNNNNNNNNNNNN NNNNNNNNNNNNNNNNNNNN   1
C  1218 hCoV-19/Kenya/NIC_087/2020|EPI_ISL_457829|2020-03    ···················· ···················· ···················· ···················· NNNNNNNNNNNNNNNNNNNN NNNNNNNNNNNNNNNNNNNN   1
C  1287 hCoV-19/England/CAMB-1AD972/2020|EPI_ISL_448029|20   ···················· ···················· ···················· ···················· NNNNNNNNNNNNNNNNNNNN NNNNNNNNNNNNNNNNNNNN   1
C  1288 hCoV-19/England/CAMB-1AD981/2020|EPI_ISL_448030|20   ···················· ···················· ···················· ···················· NNNNNNNNNNNNNNNNNNNN NNNNNNNNNNNNNNNNNNNN   1
C  1331 hCoV-19/England/CAMB-1AFF9C/2020|EPI_ISL_448091|20   ···················· ···················· ···················· ···················· NNNNNNNNNNNNNNNNNNNN NNNNNNNNNNNNNNNNNNNN   1
C  1341 hCoV-19/Scotland/EDB4383/2020|EPI_ISL_449306|2020-   ···················· ·A·················· ···················· ···················· NNNNNNNNNNNNNNNNNNNN NNNNNNNNNNNNNNNNNNNN   1
C  1394 hCoV-19/Switzerland/TI-ETHZ-100075/2020|EPI_ISL_45   ···················· ···················· ···················· ···················· ·······NNNNNNNNNNNNN NNNNNNNNNNNNNNNNNNNN   1
C  1396 hCoV-19/Switzerland/TI-ETHZ-100077/2020|EPI_ISL_45   ···················· ···················· ···················· ···················· ·······NNNNNNNNNNNNN NNNNNNNNNNNNNNNNNNNN   1
```

| | | | | | | | | | |
|---|---|---|---|---|---|---|---|---|---|
| C | 1397 | hCoV-19/Switzerland/AG-ETHZ-100080/2020\|EPI_ISL_45 | ···················· | ···················· | ···················· | ···················· | ······NNNNNNNNNNNNNN | NNNNNNNNNNNNNNNNNNNN | 1 |
| C | 1404 | hCoV-19/Switzerland/GR-ETHZ-100088/2020\|EPI_ISL_45 | ···················· | ···················· | ···················· | ···················· | ······NNNNNNNNNNNNNN | NNNNNNNNNNNNNNNNNNNN | 1 |
| C | 1405 | hCoV-19/Switzerland/GR-ETHZ-100089/2020\|EPI_ISL_45 | ···················· | ···················· | ···················· | ···················· | ······NNNNNNNNNNNNNN | ···················· | 1 |
| C | 1412 | hCoV-19/Switzerland/TI-ETHZ-100098/2020\|EPI_ISL_45 | ···················· | ···················· | ···················· | ···················· | ······NNNNNNNNNNNNNN | ···················· | 1 |
| C | 1417 | hCoV-19/Switzerland/BS-ETHZ-100111/2020\|EPI_ISL_45 | ···················· | ···················· | ···················· | ···················· | ······NNNNN··N······ | ···················· | 1 |
| C | 1418 | hCoV-19/Switzerland/BL-ETHZ-100112/2020\|EPI_ISL_45 | ···················· | ···················· | ···················· | ···················· | ······NNNNNNNNNNNNNN | ···················· | 1 |
| C | 1420 | hCoV-19/Switzerland/BL-ETHZ-100114/2020\|EPI_ISL_45 | ···················· | ···················· | ···················· | ···················· | ······NNNNNNNNNNNNNN | ···················· | 1 |
| C | 1435 | hCoV-19/England/CAMB-1B0A5B/2020\|EPI_ISL_452932\|20 | ···················· | ···················· | ···················· | ···················· | NNNNNNNNNNNNNNNNNNNN | NNNNNNNNNNNNNNNNNNNN | 1 |
| C | 1447 | hCoV-19/England/CAMB-1B19D5/2020\|EPI_ISL_452993\|20 | ···················· | ···················· | ···················· | ···················· | NNNNNNNNNNNNNNNNNNNN | NNNNNNNNNNNNNNNNNNNN | 1 |
| C | 1449 | hCoV-19/England/CAMB-1B1A2D/2020\|EPI_ISL_452996\|20 | ···················· | ···················· | ···················· | ···················· | NNNNNNNNNNNNNNNNNNNN | NNNNNNNNNNNNNNNNNNNN | 1 |
| C | 1466 | hCoV-19/Australia/VIC1593/2020\|EPI_ISL_456513\|2020 | ···················· | ···················· | ···················· | ···················· | NNNNNNNNNNNNNNNNNNNN | NNNNNNNNNNNNNNNNNNNN | 1 |
| C | 1467 | hCoV-19/Australia/VIC1604/2020\|EPI_ISL_456516\|2020 | ···················· | ···················· | ···················· | ···················· | NNNNNNNNNNNNNNNNNNNN | NNNNNNNNNNNNNNNNNNNN | 1 |
| C | 1468 | hCoV-19/Australia/VIC1607/2020\|EPI_ISL_456518\|2020 | ···················· | ···················· | ···················· | ···················· | NNNNNNNNNNNNNNNNNNNN | NNNNNNNNNNNNNNNNNNNN | 1 |
| C | 1469 | hCoV-19/Australia/VIC1608/2020\|EPI_ISL_456519\|2020 | ···················· | ···················· | ···················· | ···················· | NNNNNNNNNNNNNNNNNNNN | NNNNNNNNNNNNNNNNNNNN | 1 |
| C | 1470 | hCoV-19/Australia/VIC1609/2020\|EPI_ISL_456520\|2020 | ···················· | ···················· | ···················· | ···················· | NNNNNNNNNNNNNNNNNNNN | NNNNNNNNNNNNNNNNNNNN | 1 |
| C | 1471 | hCoV-19/Australia/VIC1611/2020\|EPI_ISL_456521\|2020 | ···················· | ···················· | ···················· | ···················· | NNNNNNNNNNNNNNNNNNNN | NNNNNNNNNNNNNNNNNNNN | 1 |
| C | 1472 | hCoV-19/Australia/VIC1618/2020\|EPI_ISL_456526\|2020 | ···················· | ···················· | ···················· | ···················· | NNNNNNNNNNNNNNNNNNNN | NNNNNNNNNNNNNNNNNNNN | 1 |
| C | 1473 | hCoV-19/Australia/VIC1621/2020\|EPI_ISL_456529\|2020 | ···················· | ···················· | ···················· | ···················· | NNNNNNNNNNNNNNNNNNNN | NNNNNNNNNNNNNNNNNNNN | 1 |
| C | 1474 | hCoV-19/Australia/VIC1623/2020\|EPI_ISL_456530\|2020 | ···················· | ···················· | ···················· | ···················· | NNNNNNNNNNNNNNNNNNNN | NNNNNNNNNNNNNNNNNNNN | 1 |
| C | 1475 | hCoV-19/Australia/VIC1624/2020\|EPI_ISL_456531\|2020 | ···················· | ···················· | ···················· | ···················· | NNNNNNNNNNNNNNNNNNNN | NNNNNNNNNNNNNNNNNNNN | 1 |
| C | 1476 | hCoV-19/Australia/VIC1639/2020\|EPI_ISL_456542\|2020 | ···················· | ···················· | ···················· | ···················· | NNNNNNNNNNNNNNNNNNNN | NNNNNNNNNNNNNNNNNNNN | 1 |
| C | 1477 | hCoV-19/Australia/VIC1640/2020\|EPI_ISL_456543\|2020 | ···················· | ···················· | ···················· | ···················· | NNNNNNNNNNNNNNNNNNNN | NNNNNNNNNNNNNNNNNNNN | 1 |
| C | 1551 | hCoV-19/Scotland/EDB4401/2020\|EPI_ISL_449309\|2020- | ···················· | ···················· | ···················· | ···················· | NNNNNNNNNNNNNNNNNNNN | NNNNNNNNNNNNNNNNNNNN | 1 |
| C | 1664 | hCoV-19/England/CAMB-1B1999/2020\|EPI_ISL_452989\|20 | ···················· | ···················· | ···················· | ···················· | NNNNNNNNNNNNNNNNNNNN | NNNNNNNNNNNNNNNNNNNN | 1 |
| C | 1734 | hCoV-19/Australia/VIC1433/2020\|EPI_ISL_456440\|2020 | ···················· | ···················· | ···················· | ···················· | NNNNNNNNNNNNNNNNNNNN | NNNNNNNNNNNNNNNNNNNN | 1 |
| C | 1735 | hCoV-19/Australia/VIC1436/2020\|EPI_ISL_456441\|2020 | ···················· | ···················· | ···················· | ···················· | NNNNNNNNNNNNNNNNNNNN | ···················· | 1 |
| C | 1736 | hCoV-19/Australia/VIC1438/2020\|EPI_ISL_456442\|2020 | ···················· | ···················· | ···················· | ···················· | NNNNNNNNNNNNNNNNNNNN | NNNNNNNNNNNNNNNNNNNN | 1 |
| C | 1737 | hCoV-19/Australia/VIC1441/2020\|EPI_ISL_456443\|2020 | ···················· | ···················· | ···················· | ···················· | NNNNNNNNNNNNNNNNNNNN | NNNNNNNNNNNNNNNNNNNN | 1 |
| C | 1739 | hCoV-19/Australia/VIC1464/2020\|EPI_ISL_456446\|2020 | ···················· | ···················· | ···················· | ···················· | NNNNNNNNNNNNNNNNNNNN | NNNNNNNNNNNNNNNNNNNN | 1 |
| C | 1740 | hCoV-19/Australia/VIC1459/2020\|EPI_ISL_456448\|2020 | ···················· | ···················· | ···················· | ···················· | NNNNNNNNNNNNNNNNNNNN | ···················· | 1 |
| C | 1741 | hCoV-19/Australia/VIC1467/2020\|EPI_ISL_456451\|2020 | ···················· | ···················· | ···················· | ···················· | NNNNNNNNNNNNNNNNNNNN | NNNN················ | 1 |
| C | 1742 | hCoV-19/Australia/VIC1458/2020\|EPI_ISL_456454\|2020 | ···················· | ···················· | ···················· | ···················· | NNNNNNNNNNNNNNNNNNNN | ···················· | 1 |
| C | 1743 | hCoV-19/Australia/VIC1489/2020\|EPI_ISL_456456\|2020 | ···················· | ···················· | ···················· | ·······S············ | NNNNNNNNNNNNNNNNNNNN | ·········R·········· | 1 |
| C | 1744 | hCoV-19/Australia/VIC1490/2020\|EPI_ISL_456457\|2020 | ···················· | ···················· | ···················· | ···················· | NNNNNNNNNNNNNNNNNNNN | NNNNNNNNNNNNNNNNNNNN | 1 |
| C | 1745 | hCoV-19/Australia/VIC1496/2020\|EPI_ISL_456460\|2020 | ···················· | ···················· | ···················· | ···················· | NNNNNNNNNNNNNNNNNNNN | NNNNNNNNNNNNNNNNNNNN | 1 |
| C | 1748 | hCoV-19/Australia/VIC1498/2020\|EPI_ISL_456464\|2020 | ···················· | ···················· | ···················· | ···················· | NNNNNNNNNNNNNNNNNNNN | ···················· | 1 |
| C | 1749 | hCoV-19/Australia/VIC1504/2020\|EPI_ISL_456468\|2020 | ···················· | ···················· | ···················· | ···················· | NNNNNNNNNNNNNNNNNNNN | NNNNNNNNNNNNNNNNNNNN | 1 |
| C | 1750 | hCoV-19/Australia/VIC1493/2020\|EPI_ISL_456471\|2020 | ···················· | ···················· | ···················· | ···················· | NNNNNNNNNNNNNNNNNNNN | NNNNNNNNNNN········· | 1 |
| C | 1751 | hCoV-19/Australia/VIC1499/2020\|EPI_ISL_456472\|2020 | ···················· | ···················· | ···················· | ···················· | NNNNNNNNNNNNNNNNNNNN | NNNNNNNNNNNNNNNNNNNN | 1 |
| C | 1752 | hCoV-19/Australia/VIC1511/2020\|EPI_ISL_456476\|2020 | ···················· | ···················· | ···················· | ···················· | NNNNNNNNNNNNNNNNNNNN | NNNNNNNNNNNNNNN····· | 1 |
| C | 1753 | hCoV-19/Australia/VIC1513/2020\|EPI_ISL_456477\|2020 | ···················· | ···················· | ···················· | ···················· | NNNNNNNNNNNNNNNNNNNN | NNNNNNNNNNNNNNNNNNNN | 1 |
| C | 1754 | hCoV-19/Australia/VIC1533/2020\|EPI_ISL_456478\|2020 | ···················· | ···················· | ···················· | ···················· | NNNNNNNNNNNNNNNNNNNN | NNNNNNNNNNNNNNNNNNNN | 1 |
| C | 1756 | hCoV-19/Australia/VIC1561/2020\|EPI_ISL_456480\|2020 | ···················· | ···················· | ···················· | ···················· | NNNNNNNNNNNNNNNNNNNN | NNNNNNNNNNNNNNNNNNNN | 1 |
| C | 1757 | hCoV-19/Australia/VIC1559/2020\|EPI_ISL_456482\|2020 | ···················· | ···················· | ···················· | ···················· | NNNNNNNNNNNNNNNNNNNN | ····M··············· | 1 |
| C | 1758 | hCoV-19/Australia/VIC1560/2020\|EPI_ISL_456483\|2020 | ···················· | ···················· | ···················· | ···················· | NNNNNNNNNNNNNNNNNNNN | NNNNNNNNNNNNNNNNNNNN | 1 |
| C | 1759 | hCoV-19/Australia/VIC1566/2020\|EPI_ISL_456484\|2020 | ···················· | ···················· | ···················· | ···················· | NNNNNNNNNNNNNNNNNNNN | NNNNNNNNNNNNNNNNNNNN | 1 |
| C | 1760 | hCoV-19/Australia/VIC1562/2020\|EPI_ISL_456492\|2020 | ···················· | ···················· | ···················· | ···················· | NNNNNNNNNNNNNNNNNNNN | ···················· | 1 |
| C | 1761 | hCoV-19/Australia/VIC1574/2020\|EPI_ISL_456493\|2020 | ···················· | ···················· | ···················· | ···················· | NNNNNNNNNNNNNNNNNNNN | NNNNNNNNNNNNNNNNNNNN | 1 |
| C | 1762 | hCoV-19/Australia/VIC1575/2020\|EPI_ISL_456495\|2020 | ···················· | ···················· | ···················· | ···················· | NNNNNNNNNNNNNNNNNNNN | NNNNNNNNNNNNNNNNNNNN | 1 |
| C | 1763 | hCoV-19/Australia/VIC1599/2020\|EPI_ISL_456499\|2020 | ···················· | ···················· | ···················· | ···················· | NNNNNNNNNNNNNNNNNNNN | NNNNNNNNNNNNNNNNNNNN | 1 |
| C | 1764 | hCoV-19/Australia/VIC1718/2020\|EPI_ISL_456619\|2020 | ···················· | ···················· | ···················· | ···················· | NNNNNNNNNNNNNNNNNNNN | NNNNNNNNNNNNNNNNNNNN | 1 |
| C | 1765 | hCoV-19/Australia/VIC1731/2020\|EPI_ISL_456630\|2020 | ···················· | ···················· | ···················· | ···················· | NNNNNNNNNNNNNNNNNNNN | NNNNNNNNNNNNNNNNNNNN | 1 |
| C | 1766 | hCoV-19/Australia/VIC1736/2020\|EPI_ISL_456633\|2020 | ···················· | ···················· | ···················· | ···················· | NNNNNNNNNNNNNNNNNNNN | NNNNNNNNNNNNNNNNNNNN | 1 |
| C | 1769 | hCoV-19/USA/MD-HP00150/2020\|EPI_ISL_457780\|2020-03 | ···················· | ···················· | ···················· | ···················· | NNNNNNNNNNNNNNNNNNNN | NNNNNNNNNNNNNNNNNNNN | 1 |
| C | 1770 | hCoV-19/USA/DC-HP00152/2020\|EPI_ISL_457781\|2020-03 | ···················· | ···················· | ···················· | ···················· | NNNNNNNNNNNNNNNNNNNN | NNNNNNNNNNNNNNNNNNNN | 1 |
| C | 1771 | hCoV-19/USA/MD-HP00153/2020\|EPI_ISL_457782\|2020-03 | ···················· | ···················· | ···················· | ···················· | NNNNNNNNNNNNNNNNNNNN | NNNNNNNNNNNNNNNNNNNN | 1 |
| C | 1773 | hCoV-19/USA/MD-HP00155/2020\|EPI_ISL_457784\|2020-03 | ···················· | ···················· | ···················· | ···················· | NNNNNNNNNNNNNNNNNNNN | NNNNNNNNNNNNNNNNNNNN | 1 |
| C | 1776 | hCoV-19/USA/MD-HP00159/2020\|EPI_ISL_457788\|2020-03 | ···················· | ···················· | ···················· | ···················· | NNNNNNNNNNNNNNNNNNNN | NNNNNNNNNNNNNNNNNNNN | 1 |
| C | 1858 | hCoV-19/Switzerland/BL-ETHZ-100837/2020\|EPI_ISL_45 | ···················· | ···················· | ···················· | ···················· | ······NNNNNNNNNNNNNN | NNNNNNNNNNNNNNNNNNNN | 1 |
| C | 1859 | hCoV-19/Switzerland/GR-ETHZ-100838/2020\|EPI_ISL_45 | ···················· | ···················· | ···················· | ···················· | ······NNNNNNNNNNNNNN | NNNNNNNNNNNNNNNNNNNN | 1 |
| C | 1862 | hCoV-19/Switzerland/AG-ETHZ-100842/2020\|EPI_ISL_45 | ···················· | ···················· | ···················· | ···················· | ······NNNNNNNNNNNNNN | NNNNNNNNNNNNNNNNNNNN | 1 |
| C | 1863 | hCoV-19/Switzerland/BL-ETHZ-100843/2020\|EPI_ISL_45 | ···············Y···· | ···················· | ···················· | ···················· | ······NNNNNNNNNNNNNN | NNNNNNNNNNNNNNNNNNNN | 1 |
| C | 1865 | hCoV-19/Switzerland/VD-ETHZ-100845/2020\|EPI_ISL_45 | ···················· | ···················· | ···················· | ···················· | ······NNNNNNNNNNNNNN | NNNNNNNNNNNNNNNNNNNN | 1 |
| C | 1866 | hCoV-19/Switzerland/VD-ETHZ-100846/2020\|EPI_ISL_45 | ···················· | ···················· | ········W··········· | ···················· | ······NNNNNNNNNNNNNN | ···················· | 1 |
| C | 1867 | hCoV-19/Switzerland/VD-ETHZ-100848/2020\|EPI_ISL_45 | ···················· | ···················A | ···················· | ···················· | ······NNNNNNNNNNNNNN | ···················· | 1 |
| C | 1892 | hCoV-19/India/OR-ILC-RMRC26/2020\|EPI_ISL_455309\|20 | ···················· | ···················· | ···················· | NNNNNNNNNNNNNNNNNNNN | ···················· | ···················· | 1 |
| C | 1928 | hCoV-19/Timor-Leste/TL25/2020\|EPI_ISL_456600\|2020- | ···················· | ···················· | ···················· | ···················· | NNNNNNNNNNNNNNNNNNNN | NNNNNNNNNNNNNNNNNNNN | 1 |
| C | 1929 | hCoV-19/Timor-Leste/TL07/2020\|EPI_ISL_456606\|2020- | ···················· | ···················· | ···················· | ···················· | NNNNNNNNNNNNNNNNNNNN | NNNNNNNNNNNNNNNNNNNN | 1 |
| C | 1930 | hCoV-19/Timor-Leste/TL21/2020\|EPI_ISL_456607\|2020- | ···················· | ···················· | ···················· | ···················· | NNNNNNNNNNNNNNNNNNNN | NNNNNNNNNNNNNNNNNNNN | 1 |
| C | 1931 | hCoV-19/Timor-Leste/TL18/2020\|EPI_ISL_456608\|2020- | ···················· | ···················· | ···················· | ···················· | NNNNNNNNNNNNNNNNNNNN | NNNNNNNNNNNNNNNNNNNN | 1 |
| C | 1932 | hCoV-19/Timor-Leste/TL23/2020\|EPI_ISL_456611\|2020- | ···················· | ···················· | ···················· | ···················· | NNNNNNNNNNNNNNNNNNNN | NNNNNNNNNNNNNNNNNNNN | 1 |
| C | 1933 | hCoV-19/Australia/VIC1711/2020\|EPI_ISL_456616\|2020 | ···················· | ···················· | ···················· | ···················· | NNNNNNNNNNNNNNNNNNNN | NNNNNNNNNNNNNNNNNNNN | 1 |
| C | 1934 | hCoV-19/Australia/VIC1712/2020\|EPI_ISL_456617\|2020 | ···················· | ···················· | ···················· | ···················· | NNNNNNNNNNNNNNNNNNNN | NNNNNNNNNNNNNNNNNNNN | 1 |
| C | 1935 | hCoV-19/Australia/VIC1719/2020\|EPI_ISL_456620\|2020 | ···················· | ···················· | ···················· | ···················· | NNNNNNNNNNNNNNNNNNNN | NNNNNNNNNNNNNNNNNNNN | 1 |
| C | 1936 | hCoV-19/Australia/VIC1769/2020\|EPI_ISL_456643\|2020 | ···················· | ·A·················· | ···················· | ···················· | NNNNNNNNNNNNNNNNNNNN | NNNNNNNNNNNNNNNNNNNN | 1 |
| C | 1937 | hCoV-19/Australia/VIC1784/2020\|EPI_ISL_456647\|2020 | ···················· | ···················· | ···················· | ···················· | NNNNNNNNNNNNNNNNNNNN | NNNNNNNNNNNNNNNNNNNN | 1 |
| C | 1938 | hCoV-19/Australia/VIC1787/2020\|EPI_ISL_456649\|2020 | ···················· | ···················· | ···················· | ···················· | NNNNNNNNNNNNNNNNNNNN | NNNNNNNNNNNNNNNNNNNN | 1 |
| C | 1939 | hCoV-19/England/BIRM-5EE63/2020\|EPI_ISL_456658\|202 | ···················· | ···················· | ···················· | ···················· | NNNNNNNNNNNNNNNNNNNN | NNNNNNNNNNNNNNNNNNNN | 1 |
| C | 1940 | hCoV-19/England/BIRM-5EE72/2020\|EPI_ISL_456659\|202 | ···················· | ···················· | ···················· | ···················· | NNNNNNNNNNNNNNNNNNNN | NNNNNNNNNNNNNNNNNNNN | 1 |
| C | 1942 | hCoV-19/England/BIRM-5EEFA/2020\|EPI_ISL_456663\|202 | ···················· | ···················· | ···················· | ···················· | NNNNNNNNNNNNNNNNNNNN | NNNNNNNNNNNNNNNNNNNN | 1 |

| | | | | | | | | |
|---|---|---|---|---|---|---|---|---|
| C | 1943 hCoV-19/England/BIRM-5EF06/2020\|EPI_ISL_456664\|202 | ···················· | ···················· | ···················· | ···················· | NNNNNNNNNNNNNNNNNNNN | NNNNNNNNNNNNNNNNNNNN | 1 |
| C | 1944 hCoV-19/England/BIRM-5EF15/2020\|EPI_ISL_456665\|202 | ···················· | ···················· | ···················· | ···················· | NNNNNNNNNNNNNNNNNNNN | NNNNNNNNNNNNNNNNNNNN | 1 |
| C | 1946 hCoV-19/England/BIRM-5F327/2020\|EPI_ISL_456671\|202 | ···················· | ···················· | ···················· | ···················· | NNNNNNNNNNNNNNNNNNNN | NNNNNNNNNNNNNNNNNNNN | 1 |
| C | 1948 hCoV-19/England/BIRM-5F372/2020\|EPI_ISL_456673\|202 | ···················· | ···················· | ···················· | ···················· | NNNNNNNNNNNNNNNNNNNN | NNNNNNNNNNNNNNNNNNNN | 1 |
| C | 2112 hCoV-19/Switzerland/BS-ETHZ-100119/2020\|EPI_ISL_45 | ···················· | ···················· | ···················· | ···················· | ·······NNNNNNNNNNNNN | NNNNNNNNNNNNNNNNNNNN | 1 |
| C | 2113 hCoV-19/Switzerland/JU-ETHZ-100120/2020\|EPI_ISL_45 | ···················· | ···················· | ···················· | ···················· | ·······NNNNNNNNNNNNN | ···················· | 1 |
| C | 2114 hCoV-19/Switzerland/TG-ETHZ-100121/2020\|EPI_ISL_45 | ···················· | ···················· | ···················· | ···················· | ·······NNNNNNNNNNNNN | ···················· | 1 |
| C | 2116 hCoV-19/Switzerland/SZ-ETHZ-100123/2020\|EPI_ISL_45 | ···················· | ···················· | ···················· | ···················· | ·······NNNNNNNNNNNNN | ···················· | 1 |
| C | 2122 hCoV-19/Switzerland/VD-ETHZ-100130/2020\|EPI_ISL_45 | ···················· | ···················· | ···················· | ···················· | ·······NNNNNNNNNNNNN | ···················· | 1 |
| C | 2125 hCoV-19/Switzerland/GR-ETHZ-100134/2020\|EPI_ISL_45 | ···················· | Y··················· | ···················· | ···················· | ·······NNNNNNNNNNNNN | NNNNNNNNNNNNNNNNNNNN | 1 |
| C | 2126 hCoV-19/Switzerland/BL-ETHZ-100135/2020\|EPI_ISL_45 | ···················· | ···················· | ···················· | ···················· | ·······NNNNNNNNNNNNN | NNNNNNNNNNNNNNNNNNNN | 1 |
| C | 2127 hCoV-19/Switzerland/BL-ETHZ-100136/2020\|EPI_ISL_45 | ···················· | ···················· | ···················· | ···················· | ·······NNNNNNNNNNNNN | NNNNNNNNNNNNNNNNNNNN | 1 |
| C | 2128 hCoV-19/Switzerland/BL-ETHZ-100137/2020\|EPI_ISL_45 | ···················· | ···················· | ···················· | ···················· | ·······NNNNNNNNNNNNN | ···················· | 1 |
| C | 2131 hCoV-19/Switzerland/SZ-ETHZ-100140/2020\|EPI_ISL_45 | ···················· | ···················· | ···················· | ···················· | ·······NNNNNNNNNNNNN | NNNNNNNNNNNNNNNNNNNN | 1 |
| C | 2133 hCoV-19/Switzerland/GR-ETHZ-100143/2020\|EPI_ISL_45 | ···················· | ···················· | ···················· | ···················· | ·······NNNNNNNNNNNNN | NNNNNNNNNNNNNNNNNNNN | 1 |
| C | 2136 hCoV-19/Switzerland/GR-ETHZ-100147/2020\|EPI_ISL_45 | ···················· | ···················· | ···················· | ···················· | ·······NNNNNNNNNNNNN | NNNNNNNNNNNNNNNNNNNN | 1 |
| C | 2142 hCoV-19/Switzerland/BL-ETHZ-100791/2020\|EPI_ISL_45 | ···················· | ···················· | ···················· | ···················· | ·······NNNNNNNNNNNNN | ···················· | 1 |
| C | 2143 hCoV-19/Switzerland/BL-ETHZ-100792/2020\|EPI_ISL_45 | ···················· | ···················· | ···················· | ···················· | ·······NNNNNNNNNNNNN | NNNNNNNNNNNNNNNNNNNN | 1 |
| C | 2146 hCoV-19/Switzerland/BE-ETHZ-100796/2020\|EPI_ISL_45 | ···················· | ···················· | ···················· | ···················· | ·······NNNNNNNNNNNNN | NNNNNNNNNNNNNNNNNNNN | 1 |
| C | 2152 hCoV-19/Switzerland/VD-ETHZ-100802/2020\|EPI_ISL_45 | ···················· | ···················· | ···················· | ···················· | ·······SNNNNNNNNNNNN | ···················· | 1 |
| C | 2153 hCoV-19/Switzerland/ZH-ETHZ-100803/2020\|EPI_ISL_45 | ···················· | ···················· | ···················· | ···················· | ·······NNNNNNNNNNNNN | ···················· | 1 |
| C | 2154 hCoV-19/Switzerland/UR-ETHZ-100804/2020\|EPI_ISL_45 | ···················· | ···················· | ···················· | ···················· | ·······NNNNNNNNNNNNN | ···················· | 1 |
| C | 2156 hCoV-19/Switzerland/BE-ETHZ-100806/2020\|EPI_ISL_45 | ···················· | ···················· | ···················· | ···················· | ·······NNNNNNNNNNNNN | NNNNNNNNNNNNNNNNNNNN | 1 |
| C | 2157 hCoV-19/Switzerland/SH-ETHZ-100808/2020\|EPI_ISL_45 | ···················· | ···················· | ···················· | ···················· | ·······NNNNNNNNNNNNN | NNNNNNNNNNNNNNNNNNNN | 1 |
| C | 2160 hCoV-19/Switzerland/ZH-ETHZ-100811/2020\|EPI_ISL_45 | ···················· | ···················· | ···················· | ···················· | ·······NNNNNNNNNNNNN | NNNNNNNNNNNNNNNNNNNN | 1 |
| C | 2163 hCoV-19/Switzerland/ZH-ETHZ-100815/2020\|EPI_ISL_45 | ···················· | ·······W············ | ···················· | ···················· | ·······NNNNNNNNNNNNN | ···················· | 1 |
| C | 2168 hCoV-19/Switzerland/BE-ETHZ-100822/2020\|EPI_ISL_45 | ···················· | ···················· | ···················· | ···················· | ·······NNNNNNNNNNNNN | NNNNNNNNNNNNNNNNNNNN | 1 |
| C | 2169 hCoV-19/Switzerland/BS-ETHZ-100823/2020\|EPI_ISL_45 | ···················· | ···················· | ···················· | ···················· | ·······NNNNNNNNNNNNN | NNNNNNNNNNNNNNNNNNNN | 1 |
| C | 2170 hCoV-19/Switzerland/TI-ETHZ-100824/2020\|EPI_ISL_45 | ···················· | ···················· | ···················· | ···················· | ·······NNNNNNNNNNNNN | ·················NN· | 1 |
| C | 2172 hCoV-19/Switzerland/SZ-ETHZ-100828/2020\|EPI_ISL_45 | ···················· | ···················· | ···················· | ···················· | ·······NNNNNNNNNNNNN | NNNNNNNNNNNNNNNNNNNN | 1 |
| C | 2173 hCoV-19/Switzerland/BS-ETHZ-100830/2020\|EPI_ISL_45 | ···················· | ···················· | ···················· | ···················· | ·······NNNNNNNNNNNNN | ···················· | 1 |
| C | 2176 hCoV-19/Switzerland/BL-ETHZ-100834/2020\|EPI_ISL_45 | ···················· | ···················· | ···················· | ···················· | ·······NNNNNNNNNNNNN | ···················· | 1 |
| C | 2179 hCoV-19/India/MH-NIV-QC-796/2020\|EPI_ISL_454567\|20 | ···················· | ···················· | ···················· | ·········NNNNNNN···· | ···················· | ···················· | 1 |
| C | 2181 hCoV-19/India/MH-NIV-QC-799/2020\|EPI_ISL_454569\|20 | ···················· | ···················· | ···················· | ·········ANNNNNNNNNN | ···················· | ···················· | 1 |
| C | 2182 hCoV-19/India/MH-NIV-QC-801/2020\|EPI_ISL_454570\|20 | ···················· | ···················· | ···················· | NNNNNNNNNNNNNNNNNNNN | ···················· | ···················· | 1 |
| C | 2247 hCoV-19/Australia/VIC1655/2020\|EPI_ISL_456557\|2020 | ···················· | ···················· | ···················· | ···················· | NNNNNNNNNNNNNNNNNNNN | NNNNNNNNNNNNNNNNNNNN | 1 |
| C | 2248 hCoV-19/Australia/VIC1657/2020\|EPI_ISL_456558\|2020 | ···················· | ···················· | ···················· | ···················· | NNNNNNNNNNNNNNNNNNNN | ·K·················· | 1 |
| C | 2249 hCoV-19/Australia/VIC1658/2020\|EPI_ISL_456559\|2020 | ···················· | ···················· | ···················· | ···················· | NNNNNNNNNNNNNNNNNNNN | NNNNNNNNNNNNNNNNNNNN | 1 |
| C | 2250 hCoV-19/Australia/VIC1659/2020\|EPI_ISL_456560\|2020 | ···················· | ···················· | ···················· | ···················· | NNNNNNNNNNNNNNNNNNNN | NNNNNNNNNNNNNNNNNNNN | 1 |
| C | 2251 hCoV-19/Australia/VIC1661/2020\|EPI_ISL_456562\|2020 | ···················· | ···················· | ···················· | ···················· | NNNNNNNNNNNNNNNNNNNN | NNNNNNNNNNNNNNNNNNNN | 1 |
| C | 2252 hCoV-19/Australia/VIC1662/2020\|EPI_ISL_456563\|2020 | ···················· | ···················· | ···················· | ···················· | NNNNNNNNNNNNNNNN···· | NNNNNNNNN··········· | 1 |
| C | 2254 hCoV-19/Australia/VIC1665/2020\|EPI_ISL_456566\|2020 | ···················· | ···················· | ···················· | ···················· | NNNNNNNNNNNNNNNNNNNN | NNNNNNNNNNNNNNNNNNNN | 1 |
| C | 2255 hCoV-19/Australia/VIC1667/2020\|EPI_ISL_456568\|2020 | ···················· | ···················· | ···················· | ···················· | NNNNNNNNNNNNNNNNNNNN | NNNNNNNNNNNNNNNNNNNN | 1 |
| C | 2256 hCoV-19/Australia/VIC1674/2020\|EPI_ISL_456573\|2020 | ···················· | ···················· | ···················· | ···················· | NNNNNNNNNNNNNNNNNNNN | NNNNNNNNNNNNNNNNNNNN | 1 |
| C | 2257 hCoV-19/Australia/VIC1675/2020\|EPI_ISL_456574\|2020 | ···················· | ···················· | ···················· | ···················· | NNNNNNNNNNNNNNNNNNNN | NNNNNNNNNNNNNNNNNNNN | 1 |
| C | 2258 hCoV-19/Australia/VIC1680/2020\|EPI_ISL_456575\|2020 | ···················· | ···················· | ···················· | ···················· | NNNNNNNNNNNNNNNNNNNN | NNNNNNNNNNNNNNNNNNNN | 1 |
| C | 2259 hCoV-19/Australia/VIC1678/2020\|EPI_ISL_456580\|2020 | ···················· | ···················· | ···················· | ···················· | NNNNNNNNNNNNNNNNNNNN | NNNNNNNNNNNNNNNNNNNN | 1 |
| C | 2260 hCoV-19/Australia/VIC1679/2020\|EPI_ISL_456581\|2020 | ···················· | ···················· | ···················· | ···················· | NNNNNNNNNNNNNNNNNNNN | NNNNNNNNNNNNNNNNNNNN | 1 |
| C | 2261 hCoV-19/Australia/VIC1704/2020\|EPI_ISL_456589\|2020 | ···················· | ···················· | ···················· | ···················· | NNNNNNNNNNNNNNNNNNNN | NNNNNNNNNNNNNNNNNNNN | 1 |
| C | 2262 hCoV-19/Australia/VIC1601/2020\|EPI_ISL_456595\|2020 | ···················· | ···················· | ···················· | ·······S············ | NNNNNNNNNNNNNNNNNNNN | NNNNNNNNNNNNNNNNNNNN | 1 |
| C | 2263 hCoV-19/Timor-Leste/TL26/2020\|EPI_ISL_456596\|2020 | ···················· | ···················· | ··K················· | ···················· | NNNNNNNNNNNNNNNNNNNN | NNNNNNNNNNNNNNNNNNNN | 1 |
| C | 2264 hCoV-19/Timor-Leste/TL05/2020\|EPI_ISL_456597\|2020- | ···················· | ···················· | ···················· | ···················· | NNNNNNNNNNNNNNNNNNNN | ···················· | 1 |
| C | 2362 hCoV-19/India/MH-NIV-12058/2020\|EPI_ISL_454524\|202 | ···················· | ···················· | ···················· | ·········NNNNNN····· | ···················· | ···················· | 1 |
| C | 2363 hCoV-19/India/MH-NIV-1738/2020\|EPI_ISL_454525\|2020 | ···················· | ···················· | ···················· | ·········NNNNNN····· | ···················· | ···················· | 1 |
| C | 2364 hCoV-19/India/MH-NIV-1804/2020\|EPI_ISL_454526\|2020 | ···················· | ···················· | ···················· | ·········NNNNNN····· | ···················· | ···················· | 1 |
| C | 2365 hCoV-19/India/MH-NIV-2387/2020\|EPI_ISL_454527\|2020 | ···················· | ···················· | ···················· | ·········NNNNNN····· | ···················· | ···················· | 1 |
| C | 2367 hCoV-19/India/MH-NIV-4130/2020\|EPI_ISL_454529\|2020 | ···················· | ···················· | ···················· | ··········NNNNNNNNNN | ···················· | ···················· | 1 |
| C | 2369 hCoV-19/India/MH-NIV-4369/2020\|EPI_ISL_454531\|2020 | ···················· | ···················· | ···················· | ·········NNNNNNNNNNN | ···················· | ···················· | 1 |
| C | 2370 hCoV-19/India/MH-NIV-4379/2020\|EPI_ISL_454532\|2020 | ···················· | ···················· | ···················· | ····TNNNNNNNNNNNNNNN | ···················· | ···················· | 1 |
| C | 2373 hCoV-19/India/MH-NIV-5102/2020\|EPI_ISL_454536\|2020 | ···················· | ···················· | ···················· | ·········NNNNNNNNNNN | ···················· | ···················· | 1 |
| C | 2374 hCoV-19/India/MH-NIV-5104/2020\|EPI_ISL_454537\|2020 | ···················· | ···················· | ···················· | NNNNNNNNNNNNNNNNNNNN | ···················· | ···················· | 1 |
| C | 2377 hCoV-19/India/MH-NIV-6423/2020\|EPI_ISL_454543\|2020 | ···················· | ···················· | ···················· | ·········NNNNNN····· | ···················· | ···················· | 1 |
| C | 2382 hCoV-19/India/MH-NIV-7893/2020\|EPI_ISL_454551\|2020 | ···················· | ···················· | ···················· | ·········NNNNNN····· | ···················· | ···················· | 1 |
| C | 2383 hCoV-19/India/MH-NIV-7966/2020\|EPI_ISL_454552\|2020 | ···················· | ···················· | ···················· | NNNNNNNNNNNNNNNNNNNN | ···················· | ···················· | 1 |
| C | 2384 hCoV-19/India/MH-NIV-9154/2020\|EPI_ISL_454557\|2020 | ···················· | ···················· | ···················· | ·········NNNNNN····· | ···················· | ···················· | 1 |
| C | 2386 hCoV-19/India/MH-NIV-701-3/2020\|EPI_ISL_454560\|202 | ···················· | ···················· | ···················· | ·········TNNNNNNNNNN | ···················· | ···················· | 1 |
| C | 2390 hCoV-19/India/MH-NIV-QA-708/2020\|EPI_ISL_454564\|20 | ···················· | ···················· | ···················· | NNNNNNNNNNNNNNNNNNNN | ···················· | ···················· | 1 |
| C | 2391 hCoV-19/India/MH-NIV-QA-710/2020\|EPI_ISL_454565\|20 | ···················· | ···················· | ···················· | ·········NNNNNN····· | ···················· | ···················· | 1 |
| C | 2539 hCoV-19/USA/MA-MGH-00532/2020\|EPI_ISL_460381\|2020- | ···················· | ···················· | ···················· | ···················· | NNNNNNNNNNNNNNNNNNNN | ···················· | 1 |
| C | 2560 hCoV-19/Scotland/CVR3076/2020\|EPI_ISL_461660\|2020- | ···················· | ·A·················· | ···················· | ···················· | NNNNNNNNNNNNNNNNNNNN | ···················· | 1 |
| C | 2573 hCoV-19/Israel/2113255/2020\|EPI_ISL_447344\|2020-04 | ···················· | ···················· | ···················· | ···················· | ·······NNNNNNNNNNNNN | ···················· | 1 |
| C | 2648 hCoV-19/Thailand/Bangkok-CONI-0095/2020\|EPI_ISL_45 | ···················· | ···················· | ···················· | ···················· | NNNNNNNNNNNN········ | ···················· | 1 |
| C | 2751 hCoV-19/USA/MA-MGH-00091/2020\|EPI_ISL_460348\|2020- | ···················· | ···················· | ···················· | ···················· | ·············NNNNNNN | ···················· | 1 |
| C | 2777 hCoV-19/Scotland/CVR3016/2020\|EPI_ISL_461624\|2020- | ···················· | ···················A | ···················· | ···················· | NNNNNNNNNNNNNNNNNNNN | NNNNNNNNNNNNNNNNNNNN | 1 |
| C | 2778 hCoV-19/Scotland/CVR3032/2020\|EPI_ISL_461625\|2020- | ···················· | ···················· | ···················· | ···················· | NNNNNNNNNNNNNNNNNNNN | NNNNNNNNNNNNNNNNNNNN | 1 |
| C | 2782 hCoV-19/Scotland/CVR3042/2020\|EPI_ISL_461630\|2020- | ···················· | ···················· | ···················· | ···················· | NNNNNNNNNNNNNNNNNNNN | NNNNNNNNNNNNNNNNNNNN | 1 |
| C | 2784 hCoV-19/Scotland/CVR3045/2020\|EPI_ISL_461633\|2020- | ···················· | ···················· | ···················· | ···················· | NNNNNNNNNNNNNNNNNNNN | ···················· | 1 |
| C | 2796 hCoV-19/Scotland/CVR3078/2020\|EPI_ISL_461662\|2020- | ···················· | ···················· | ···················· | ···················· | NNNNNNNNNNNNNNNNNNNN | ···················· | 1 |
| C | 2799 hCoV-19/Scotland/CVR3084/2020\|EPI_ISL_461667\|2020- | ···················· | ···················· | ···················· | ···················· | NNNNNNNNNNNNNNNNNNNN | ···················· | 1 |
| C | 2803 hCoV-19/Scotland/CVR3101/2020\|EPI_ISL_461674\|2020- | ···················· | ·A·················· | ···················· | ···················· | NNNNNNNNNNNNNNNNNNNN | ···················· | 1 |

| | | | | | | | | |
|---|---|---|---|---|---|---|---|---|
| C | 2808 hCoV-19/Scotland/CVR3116/2020\|EPI_ISL_461688\|2020- | ···················· | ···················· | ···················· | ···················· | NNNNNNNNNNNNNNNNNNNN | ···················· | 1 |
| C | 2991 hCoV-19/Scotland/EDB4881/2020\|EPI_ISL_453169\|2020- | ···················· | ···················· | ···················· | ···················· | NNNNNNNNNNNNNNNNNNNN | NNNNNNNNNNNNNNNNNNNN | 1 |
| C | 2992 hCoV-19/Scotland/EDB4882/2020\|EPI_ISL_453170\|2020- | ···················· | ·A·················· | ···················· | ···················· | NNNNNNNNNNNNNNNNNNNN | NNNNNNNNNNNNNNNNNNNN | 1 |
| C | 2993 hCoV-19/Scotland/EDB4883/2020\|EPI_ISL_453171\|2020- | ···················· | ·A·················· | ···················· | ···················· | NNNNNNNNNNNNNNNNNNNN | NNNNNNNNNNNNNNNNNNNN | 1 |
| C | 2995 hCoV-19/Scotland/EDB4887/2020\|EPI_ISL_453173\|2020- | ···················· | ·A·················· | ···················· | ···················· | NNNNNNNNNNNNNNNNNNNN | NNNNNNNNNNNNNNNNNNNN | 1 |
| C | 2996 hCoV-19/Scotland/EDB4890/2020\|EPI_ISL_453174\|2020- | ···················· | ·A·················· | ···················· | ···················· | NNNNNNNNNNNNNNNNNNNN | NNNNNNNNNNNNNNNNNNNN | 1 |
| C | 3001 hCoV-19/Scotland/EDB5017/2020\|EPI_ISL_453181\|2020- | ···················· | ···················· | ···················· | ···················· | NNNNNNNNNNNNNNNNNNNN | NNNNNNNNNNNNNNNNNNNN | 1 |
| C | 3070 hCoV-19/Scotland/CVR3008/2020\|EPI_ISL_461617\|2020- | ···················· | ···················A | ···················· | ···················· | NNNNNNNNNNNNNNNNNNNN | NNNNNNNNNNNNNNNNNNNN | 1 |
| C | 3076 hCoV-19/Israel/130711104/2020\|EPI_ISL_447455\|2020- | ···················· | ···················· | ···················· | ···················· | NNNNNNNNNNNNNNNNNNNN | ·················N·N | 1 |
| C | 3079 hCoV-19/Spain/VC-FISABIO-471/2020\|EPI_ISL_447471\|2 | ···················· | ···················· | ···················· | ···················· | ·NNNNNNNNNNNNNNNNNNN | ···················· | 1 |
| C | 3086 hCoV-19/Spain/VC-FISABIO-537/2020\|EPI_ISL_447482\|2 | ···················· | ···················· | ···················· | ···················· | NNNNNNNNNNNNNNNNNNNN | ···················· | 1 |
| C | 3087 hCoV-19/Spain/VC-FISABIO-538/2020\|EPI_ISL_447483\|2 | ···················· | ···················· | ···················· | ···················· | NNNNNNNNNNNNNNNNNNNN | NNNNNNNNNNNNNNNNNNNN | 1 |
| C | 3088 hCoV-19/Spain/VC-FISABIO-540/2020\|EPI_ISL_447485\|2 | ···················· | ···················· | ···················· | ···················· | ···················· | NNNNNNNNNNNNNNN····· | 1 |
| C | 3211 hCoV-19/Thailand/Bangkok-CONI-0109/2020\|EPI_ISL_45 | ···················· | ···················· | ···················· | ···················· | NNNNNNNNNNNNNNNNNNNN | ···················· | 1 |
| C | 3213 hCoV-19/Thailand/Bangkok-CONI-0113/2020\|EPI_ISL_45 | ···················· | ···················· | ···················· | ···················· | NNNNNNNNNNNN········ | ···················· | 1 |
| C | 3230 hCoV-19/Thailand/Bangkok-CONI-0149/2020\|EPI_ISL_45 | ···················· | ···················· | ···················· | ···················· | ·NNNNNNNNNNNN······· | ···················· | 1 |
| C | 3329 hCoV-19/Israel/2046815/2020\|EPI_ISL_447400\|2020-04 | ···················· | ···················· | ···················· | ···················· | ···················· | NN···N····N·NNN·NNNN | 1 |
| C | 3337 hCoV-19/Israel/13077562/2020\|EPI_ISL_447430\|2020-0 | ···················· | ······N·N····NNN·N·· | ···················· | ···················· | ···················· | ···················· | 1 |
| C | 3362 hCoV-19/Israel/130710062/2020\|EPI_ISL_447469\|2020- | ···················· | ···················· | ···················· | ···················· | ······N··NNNNNNNNNNN | NNNNNNNNNNNNNNNNNNNN | 1 |
| C | 3569 hCoV-19/Scotland/EDB5470/2020\|EPI_ISL_461745\|2020- | ···················· | ···················· | ···················· | ···················· | NNNNNNNNNNNNNNNNNNNN | NNNNNNNNNNNNNNNNNNNN | 1 |
| C | 3570 hCoV-19/Scotland/EDB5472/2020\|EPI_ISL_461746\|2020- | ···················· | ···················· | ···················· | ···················· | NNNNNNNNNNNNNNNNNNNN | NNNNNNNNNNNNNNNNNNNN | 1 |
| C | 3621 hCoV-19/England/CAMB-1B2093/2020\|EPI_ISL_453003\|20 | ···················· | ···················· | ···················· | ···················· | NNNNNNNNNNNNNNNNNNNN | NNNNNNNNNNNNNNNNNNNN | 1 |
| C | 3629 hCoV-19/Scotland/CVR2257/2020\|EPI_ISL_453034\|2020- | ···················· | ···················· | ···················· | ···················· | NNNNNNNNNNNNNNNNNNNN | NNNNNNNNNNNNNNNNNNNN | 1 |
| C | 3780 hCoV-19/Scotland/EDB3021/2020\|EPI_ISL_456909\|2020- | ···················· | ···················· | ···················· | ···················· | NNNNNNNNNNNNNNNNNNNN | NNNNNNNNNNNNNNNNNNNN | 1 |
| C | 3807 hCoV-19/Scotland/EDB5244/2020\|EPI_ISL_456991\|2020- | ···················· | ···················· | ···················· | ···················· | NNNNNNNNNNNNNNNNNNNN | NNNNNNNNNNNNNNNNNNNN | 1 |
| C | 3809 hCoV-19/Scotland/EDB5247/2020\|EPI_ISL_456993\|2020- | ···················· | ·A·················· | ···················· | ···················· | NNNNNNNNNNNNNNNNNNNN | NNNNNNNNNNNNNNNNNNNN | 1 |
| C | 3811 hCoV-19/Scotland/EDB5250/2020\|EPI_ISL_456995\|2020- | ···················· | ···················· | ···················· | ···················· | NNNNNNNNNNNNNNNNNNNN | NNNNNNNNNNNNNNNNNNNN | 1 |
| C | 3815 hCoV-19/Scotland/EDB5288/2020\|EPI_ISL_456999\|2020- | ···················· | ···················· | ···················· | ···················· | NNNNNNNNNNNNNNNNNNNN | NNNNNNNNNNNNNNNNNNNN | 1 |
| C | 3846 hCoV-19/England/NOTT-110EB7/2020\|EPI_ISL_448432\|20 | ···················· | ···················· | ···················· | ···················· | NNNNNNNNNNNNNNNNNNNN | NNNNNNNNNNNNNNNNNNNN | 1 |
| C | 3933 hCoV-19/Scotland/EDB3369/2020\|EPI_ISL_456915\|2020- | ···················· | ···················· | ···················· | ···················· | NNNNNNNNNNNNNNNNNNNN | NNNNNNNNNNNNNNNNNNNN | 1 |
| C | 3935 hCoV-19/Scotland/EDB4103/2020\|EPI_ISL_456918\|2020- | ···················· | ···················A | ···················· | ···················· | NNNNNNNNNNNNNNNNNNNN | NNNNNNNNNNNNNNNNNNNN | 1 |
| C | 3952 hCoV-19/Scotland/EDB4477/2020\|EPI_ISL_456941\|2020- | ···················· | ·A·················· | ···················· | ···················· | ·······N·····NN····· | ·······NN·NN·NN····· | 1 |
| C | 3953 hCoV-19/Scotland/EDB4479/2020\|EPI_ISL_456942\|2020- | ···················· | ·A·················· | ···················· | ···················· | ··NN·····NNNNNNN···· | NNNN·N·N·····NN····· | 1 |
| C | 3957 hCoV-19/Scotland/EDB4507/2020\|EPI_ISL_456946\|2020- | ···················· | ···················· | ···················· | ···················· | NNNNNNNNNNNNNNNNNNNN | NNNNNNNNNNNNNNNNNNNN | 1 |
| C | 3958 hCoV-19/Scotland/EDB4508/2020\|EPI_ISL_456947\|2020- | ···················· | ···················· | ···················· | ···················· | NNNNNNNNNNNNNNNNNNNN | NNNNNNNNNNNNNNNNNNNN | 1 |
| C | 3961 hCoV-19/Scotland/EDB5039/2020\|EPI_ISL_456951\|2020- | ···················· | ···················· | ···················· | ···················· | NNNNNNNNNNNNNNNNNNNN | NNNNNNNNNNNNNNNNNNNN | 1 |
| C | 4019 hCoV-19/USA/MA-MGH-00567/2020\|EPI_ISL_460166\|2020- | ·······NNNNNNNNNNNNN | ···················· | ···················· | ···················· | ···················· | ···················· | 1 |
| C | 4103 hCoV-19/England/NOTT-110CF9/2020\|EPI_ISL_448407\|20 | ···················· | ···················· | ···················· | ···················· | NNNNNNNNNNNNNNNNNNNN | NNNNNNNNNNNNNNNNNNNN | 1 |
| C | 4401 hCoV-19/Scotland/EDB4236/2020\|EPI_ISL_453117\|2020- | ···················· | ···················A | ···················· | ···················· | NNNNNNNNNNNNNNNNNNNN | NNNNNNNNNNNNNNNNNNNN | 1 |
| C | 4420 hCoV-19/Scotland/EDB4755/2020\|EPI_ISL_453148\|2020- | ···················· | ·A·················· | ···················· | ···················· | NNNNNNNNNNNNNNNNNNNN | NNNNNNNNNNNNNNNNNNNN | 1 |
| C | 4430 hCoV-19/Scotland/EDB4834/2020\|EPI_ISL_453160\|2020- | ···················· | ···················· | ···················· | ···················· | NNNNNNNNNNNNNNNNNNNN | NNNNNNNNNNNNNNNNNNNN | 1 |
| C | 4520 hCoV-19/Spain/VC-FISABIO-654/2020\|EPI_ISL_455743\|2 | ···················· | ···················· | ···················· | ···················· | NNNNNNNNNNNNNNNNNNNN | NNNNNNNNNNNNNNNNNNNN | 1 |
| C | 4603 hCoV-19/Scotland/CVR2925/2020\|EPI_ISL_461592\|2020- | ···················· | ···················· | ···················· | ···················· | NNNNNNNNNNNNNNNNNNNN | NNNNNNNNNNNNNNNNNNNN | 1 |
| C | 4608 hCoV-19/Scotland/CVR2938/2020\|EPI_ISL_461599\|2020- | ···················· | ···················· | ···················· | ···················· | NNNNNNNNNNNNNNNNNNNN | ···················· | 1 |
| C | 4632 hCoV-19/Israel/701002752/2020\|EPI_ISL_447284\|2020- | ···················· | ···················· | ···················· | ···················· | ·········NNNNNN···NN | ·········NN········· | 1 |
| C | 4741 hCoV-19/Spain/VC-FISABIO-656/2020\|EPI_ISL_455730\|2 | ···················· | ···················· | ···················· | ···················· | NNNNNNNNNNNNNNNNNNNN | NNNNNNNNNNNNNNNNNNNN | 1 |
| C | 4743 hCoV-19/Spain/VC-FISABIO-652/2020\|EPI_ISL_455742\|2 | ···················· | ···················· | ···················· | ···················· | NNNNNNNNNNNNNNNNNNNN | NNNNNNNNNNNNNNNNNNNN | 1 |
| C | 4817 hCoV-19/England/BIRM-5F4D9/2020\|EPI_ISL_461508\|202 | ···················· | ···················· | ···················· | ···················· | NNNNNNNNNNNNNNNNNNNN | NNNNNNNNNNNNNNNNNNNN | 1 |
| C | 4818 hCoV-19/England/BIRM-5F4E8/2020\|EPI_ISL_461509\|202 | ···················· | ···················· | ···················· | ···················· | NNNNNNNNNNNNNNNNNNNN | NNNNNNNNNNNNNNNNNNNN | 1 |
| C | 4819 hCoV-19/England/BIRM-5F4F7/2020\|EPI_ISL_461510\|202 | ···················· | ···················· | ···················· | ···················· | NNNNNNNNNNNNNNNNNNNN | NNNNNNNNNNNNNNNNNNNN | 1 |
| C | 4821 hCoV-19/England/BIRM-5F54F/2020\|EPI_ISL_461514\|202 | ···················· | ···················· | ···················· | ···················· | NNNNNNNNNNNNNNNNNNNN | NNNNNNNNNNNNNNNNNNNN | 1 |
| C | 4822 hCoV-19/England/BIRM-5F56D/2020\|EPI_ISL_461515\|202 | ···················· | ···················· | ···················· | ···················· | NNNNNNNNNNNNNNNNNNNN | NNNNNNNNNNNNNNNNNNNN | 1 |
| C | 4823 hCoV-19/England/BIRM-5F5A9/2020\|EPI_ISL_461517\|202 | ···················· | ···················· | ···················· | ···················· | NNNNNNNNNNNNNNNNNNNN | NNNNNNNNNNNNNNNNNNNN | 1 |
| C | 4825 hCoV-19/England/BIRM-5F61F/2020\|EPI_ISL_461520\|202 | ···················· | ···················· | ···················· | ···················· | NNNNNNNNNNNNNNNNNNNN | NNNNNNNNNNNNNNNNNNNN | 1 |
| C | 4826 hCoV-19/England/BIRM-5F63D/2020\|EPI_ISL_461521\|202 | ···················· | ···················· | ···················· | ···················· | NNNNNNNNNNNNNNNNNNNN | NNNNNNNNNNNNNNNNNNNN | 1 |
| C | 4828 hCoV-19/England/BIRM-5F697/2020\|EPI_ISL_461524\|202 | ···················· | ···················· | ···················· | ···················· | NNNNNNNNNNNNNNNNNNNN | NNNNNNNNNNNNNNNNNNNN | 1 |
| C | 4829 hCoV-19/England/BIRM-5F6A6/2020\|EPI_ISL_461525\|202 | ···················· | ···················· | ···················· | ···················· | NNNNNNNNNNNNNNNNNNNN | NNNNNNNNNNNNNNNNNNNN | 1 |
| C | 4830 hCoV-19/England/BIRM-5FDCF/2020\|EPI_ISL_461526\|202 | ···················· | ···················· | ···················· | ···················· | NNNNNNNNNNNNNNNNNNNN | NNNNNNNNNNNNNNNNNNNN | 1 |
| C | 4831 hCoV-19/England/BIRM-5FE35/2020\|EPI_ISL_461527\|202 | ···················· | ···················· | ···················· | ···················· | NNNNNNNNNNNNNNNNNNNN | NNNNNNNNNNNNNNNNNNNN | 1 |
| C | 4834 hCoV-19/England/BIRM-5FE80/2020\|EPI_ISL_461530\|202 | ···················· | ···················· | ···················· | ···················· | NNNNNNNNNNNNNNNNNNNN | NNNNNNNNNNNNNNNNNNNN | 1 |
| C | 4835 hCoV-19/England/BIRM-5FE9F/2020\|EPI_ISL_461531\|202 | ···················· | ···················· | ···················· | ···················· | NNNNNNNNNNNNNNNNNNNN | NNNNNNNNNNNNNNNNNNNN | 1 |
| C | 4837 hCoV-19/England/BIRM-5FEDB/2020\|EPI_ISL_461534\|202 | ···················· | ···················· | ···················· | ···················· | NNNNNNNNNNNNNNNNNNNN | NNNNNNNNNNNNNNNNNNNN | 1 |
| C | 4841 hCoV-19/England/BIRM-5FF7E/2020\|EPI_ISL_461541\|202 | ···················· | ···················· | ···················· | ···················· | NNNNNNNNNNNNNNNNNNNN | NNNNNNNNNNNNNNNNNNNN | 1 |
| C | 4842 hCoV-19/England/BIRM-5FF8D/2020\|EPI_ISL_461542\|202 | ···················· | ···················· | ···················· | ···················· | NNNNNNNNNNNNNNNNNNNN | NNNNNNNNNNNNNNNNNNNN | 1 |
| C | 4844 hCoV-19/England/BIRM-5FFAB/2020\|EPI_ISL_461544\|202 | ···················· | ···················· | ···················· | ···················· | NNNNNNNNNNNNNNNNNNNN | NNNNNNNNNNNNNNNNNNNN | 1 |
| C | 4845 hCoV-19/England/BIRM-5FFBA/2020\|EPI_ISL_461545\|202 | ···················· | ···················· | ···················· | ···················· | NNNNNNNNNNNNNNNNNNNN | NNNNNNNNNNNNNNNNNNNN | 1 |
| C | 4847 hCoV-19/England/CAMB-1B3074/2020\|EPI_ISL_461547\|20 | ···················· | ···················· | ···················· | ···················· | NNNNNNNNNNNNNNNNNNNN | NNNNNNNNNNNNNNNNNNNN | 1 |
| C | 4848 hCoV-19/England/CAMB-1B3083/2020\|EPI_ISL_461548\|20 | ···················· | ···················· | ···················· | ···················· | NNNNNNNNNNNNNNNNNNNN | NNNNNNNNNNNNNNNNNNNN | 1 |
| C | 4849 hCoV-19/England/CAMB-1B30ED/2020\|EPI_ISL_461552\|20 | ···················· | ···················· | ···················· | ···················· | NNNNNNNNNNNNNNNNNNNN | NNNNNNNNNNNNNNNNNNNN | 1 |
| C | 5043 hCoV-19/Spain/AN-IBV-002063/2020\|EPI_ISL_452367\|20 | ···················· | ···················· | ···················· | ···················· | NNNNNNNNNNNNNNNNNNNN | NNNNNNNN···NNNN····· | 1 |
| C | 5046 hCoV-19/Spain/AN-IBV-002055/2020\|EPI_ISL_452374\|20 | ···················· | ···················· | ···················· | ···················· | NNNNNNNNNNNNNNNNNNNN | NNNNNNNNNNNNNNNNNNNN | 1 |
| C | 5047 hCoV-19/Spain/AN-IBV-002048/2020\|EPI_ISL_452379\|20 | ···················· | ···················· | ···················· | ···················· | NNNNNNNNNNNNNNNNNNNN | NNNNNNNNNNNNNNNNNNNN | 1 |
| C | 5048 hCoV-19/Spain/AN-IBV-002047/2020\|EPI_ISL_452380\|20 | ···················· | ···················· | ···················· | ···················· | NNNNNNNNNNNNNNNNNNNN | NNNNNNNNNNNNNNNNNNNN | 1 |
| C | 5049 hCoV-19/Spain/AN-IBV-002046/2020\|EPI_ISL_452381\|20 | ···················· | ···················· | ···················· | ···················· | NNNNNNNNNNNNNNNNNNNN | NNNNNNNNNNNNNNNNNNNN | 1 |
| C | 5050 hCoV-19/Spain/AN-IBV-002043/2020\|EPI_ISL_452383\|20 | ···················· | ···················· | ···················· | ···················· | ··NNNNN······N······ | NNN················· | 1 |
| C | 5051 hCoV-19/Spain/AN-IBV-002041/2020\|EPI_ISL_452385\|20 | ···················· | ···················· | ···················· | ···················· | NNNNNNNNNNNNNNNNNNNN | ···················· | 1 |
| C | 5052 hCoV-19/Spain/AN-IBV-002038/2020\|EPI_ISL_452387\|20 | ···················· | ···················· | ···················· | ···················· | NNNNNNNNNNNNNNNNNNNN | NNNNNNNNNNNNNNN····· | 1 |
| C | 5053 hCoV-19/Spain/AN-IBV-002027/2020\|EPI_ISL_452392\|20 | ···················· | ···················· | ···················· | ···················· | NNNNNNNNNNNNNNNNNNNN | NNNNNNNNNNNNNNNNNNNN | 1 |
| C | 5054 hCoV-19/Spain/AN-IBV-002025/2020\|EPI_ISL_452394\|20 | ···················· | ···················· | ···················· | ···················· | NNNNNNNNNNNNNNNNNNNN | NNNNN··············· | 1 |
| C | 5055 hCoV-19/Spain/AN-IBV-002003/2020\|EPI_ISL_452396\|20 | ···················· | ···················· | ···················· | ···················· | NNNNNNNNNNNNNNNNNNNN | NNNNNNNNNNNNNNNNNNNN | 1 |

| | | | | | | | | |
|---|---|---|---|---|---|---|---|---|
| C | 5056 hCoV-19/Spain/AN-IBV-002001/2020\|EPI_ISL_452397\|20 | ···················· | ···················· | ···················· | ···················· | NNNNNNNNNNNNNNNNNNNN | NNNNNNNNNNNNNNNNNNNN | 1 |
| C | 5057 hCoV-19/Spain/AN-IBV-002000/2020\|EPI_ISL_452398\|20 | ···················· | ···················· | ···················· | ···················· | ···················· | NNNNNNNNNNNNNNN····· | 1 |
| C | 5058 hCoV-19/Scotland/CVR2472/2020\|EPI_ISL_453052\|2020- | ···················· | ···················A | ···················· | ···················· | NNNNNNNNNNNNNNNNNNNN | NNNNNNNNNNNNNNNNNNNN | 1 |
| C | 5072 hCoV-19/Scotland/CVR2882/2020\|EPI_ISL_453081\|2020- | ···················· | ···················· | ···················· | ···················· | NNNNNNNNNNNNNNNNNNNN | ···················· | 1 |
| C | 5079 hCoV-19/Scotland/CVR3188/2020\|EPI_ISL_453097\|2020- | ···················· | ·A·················· | ···················· | ···················· | NNNNNNNNNNNNNNNNNNNN | ···················· | 1 |
| C | 5187 hCoV-19/England/NORW-E96DE/2020\|EPI_ISL_453612\|202 | ···················· | ···················· | ···················· | ···················· | NNNNNNNNNNNNNNNNNNNN | NNNNNNNNN··········· | 1 |
| C | 5207 hCoV-19/England/NOTT-111153/2020\|EPI_ISL_453635\|20 | ···················· | ···················· | ···················· | ···················· | NNNNNNNNNNNNNNNNNNNN | NNNNNNNNNNNNNNNNNNNN | 1 |
| C | 5405 hCoV-19/Colombia/CUN-GVI-93301/2020\|EPI_ISL_447769 | ···················· | ···················· | ···················· | ···················· | ···················· | NNNNNNNNNNNNNNNNNNNN | 1 |
| C | 5491 hCoV-19/England/EXET-135E52/2020\|EPI_ISL_457173\|20 | ···················· | ···················· | ···················· | ···················· | NNNNNNNNNNNNNNNNNNNN | NNNNNNNNNNNNNNNNNNNN | 1 |
| C | 5502 hCoV-19/England/EXET-13605B/2020\|EPI_ISL_457191\|20 | ···················· | ···················· | ···················· | ···················· | NNNNNNNNNNNNNNNNNNNN | NNNNNNNNNNNNNNNNNNNN | 1 |
| C | 5504 hCoV-19/England/EXET-136097/2020\|EPI_ISL_457194\|20 | ···················· | ···················· | ···················· | ···················· | NNNNNNNNNNNNNNNNNNNN | NNNNNNNNNNNNNNNNNNNN | 1 |
| C | 5506 hCoV-19/England/EXET-1360C4/2020\|EPI_ISL_457196\|20 | ···················· | ···················· | ···················· | ···················· | NNNNNNNNNNNNNNNNNNNN | NNNNNNNNNNNNNNNNNNNN | 1 |
| C | 5650 hCoV-19/England/BIRM-5EBB7/2020\|EPI_ISL_447931\|202 | ···················· | ···················· | ···················· | ···················· | NNNNNNNNNNNNNNNNNNNN | NNNNNNNNNNNNNNNNNNNN | 1 |
| C | 5651 hCoV-19/England/BIRM-5EC1E/2020\|EPI_ISL_447933\|202 | ···················· | ···················· | ···················· | ···················· | NNNNNNNNNNNNNNNNNNNN | NNNNNNNNNNNNNNNNNNNN | 1 |
| C | 5652 hCoV-19/England/BIRM-5EC5A/2020\|EPI_ISL_447936\|202 | ···················· | ···················· | ···················· | ···················· | NNNNNNNNNNNNNNNNNNNN | NNNNNNNNNNNNNNNNNNNN | 1 |
| C | 5654 hCoV-19/England/BIRM-5ED48/2020\|EPI_ISL_447940\|202 | ···················· | ···················· | ···················· | ···················· | NNNNNNNNNNNNNNNNNNNN | NNNNNNNNNNNNNNNNNNNN | 1 |
| C | 5730 hCoV-19/Spain/AN-IBV-001993/2020\|EPI_ISL_452400\|20 | ···················· | ···················· | ···················· | ···················· | ·NNNNNN············· | ···················· | 1 |
| C | 5731 hCoV-19/Spain/AN-IBV-001988/2020\|EPI_ISL_452402\|20 | ···················· | ···················· | ···················· | ···················· | NNNNNNNNNNNNNNNNNNNN | NNNNNNNNNNNNNNNNNNNN | 1 |
| C | 5732 hCoV-19/Spain/AN-IBV-001982/2020\|EPI_ISL_452404\|20 | ···················· | ···················· | ···················· | ···················· | NNNNNNNNNNNNNNNNNNNN | ···················· | 1 |
| C | 5733 hCoV-19/Spain/AN-IBV-001979/2020\|EPI_ISL_452405\|20 | ···················· | ···················· | ···················· | ···················· | NNNNNNNNNNNNNNNNNNNN | ···NNNNNNNNNNNNNNNNN | 1 |
| C | 5734 hCoV-19/Spain/AN-IBV-001978/2020\|EPI_ISL_452406\|20 | ···················· | ···················· | ···················· | ···················· | NNNNNNNNNNNNNNNNNNNN | ···················· | 1 |
| C | 5735 hCoV-19/Spain/AN-IBV-001977/2020\|EPI_ISL_452407\|20 | ···················· | ···················· | ···················· | ···················· | NNNNNNNNNNNNNNNNNNNN | NNNNNNNNNNNNNNNNNNNN | 1 |
| C | 5736 hCoV-19/Spain/AN-IBV-001973/2020\|EPI_ISL_452409\|20 | ···················· | ···················· | ···················· | ···················· | NNNNNNNNNNNNNNNNNNNN | NNNNNNNNNNNNNNNNNNNN | 1 |
| C | 5737 hCoV-19/Spain/AN-IBV-001972/2020\|EPI_ISL_452410\|20 | ···················· | ···················· | ···················· | ···················· | NNNNNNNNNNNNNNNNNNNN | NNNNNNNNNNNNNNN····N | 1 |
| C | 5738 hCoV-19/Spain/AN-IBV-001971/2020\|EPI_ISL_452411\|20 | ···················· | ···················· | ···················· | ···················· | NNNNNNN············· | NNNNNNNNNNNNNNNNNNNN | 1 |
| C | 5740 hCoV-19/Spain/AN-IBV-001968/2020\|EPI_ISL_452414\|20 | ···················· | ···················· | ···················· | ···················· | NNNNNNN············· | ···················· | 1 |
| C | 5741 hCoV-19/Spain/AN-IBV-001953/2020\|EPI_ISL_452426\|20 | ···················· | ···················· | ···················· | ···················· | NNNNNNNNNNNNN······· | ···················· | 1 |
| C | 5742 hCoV-19/Spain/AN-IBV-001950/2020\|EPI_ISL_452429\|20 | ···················· | ···················· | ···················· | ···················· | NNNNNNNNNNNNNNNNNNNN | NNNNNNNNNNNNNNN····· | 1 |
| C | 5743 hCoV-19/Spain/AN-IBV-001949/2020\|EPI_ISL_452430\|20 | ···················· | ···················· | ···················· | ···················· | NNNNNNNNNNNNNNNNNNNN | NNNNNNNNNNNNNNNNNNNN | 1 |
| C | 5744 hCoV-19/Spain/AN-IBV-001947/2020\|EPI_ISL_452432\|20 | ···················· | ···················· | ···················· | ···················· | NNNNNNNNNNNNNNNNNNNN | NNNNNNNNNNNNNNNNNNNN | 1 |
| C | 5745 hCoV-19/Spain/AN-IBV-001946/2020\|EPI_ISL_452433\|20 | ···················· | ···················· | ···················· | ···················· | NNNNNNNNNNNNNNNNNNNN | NNNNNNNNNNNNNNNNNNNN | 1 |
| C | 5747 hCoV-19/Spain/VC-IBV-001930/2020\|EPI_ISL_452444\|20 | ···················· | ···················· | ···················· | ···················· | NNNNNNNNNNNNNNNNNNNN | ···················· | 1 |
| C | 5748 hCoV-19/Spain/AN-IBV-001927/2020\|EPI_ISL_452447\|20 | ···················· | ···················· | ···················· | ···················· | NNNNNNNNNNNNNNNNNNNN | NNNNNNNNNNNNNNNNNNNN | 1 |
| C | 5749 hCoV-19/Spain/AN-IBV-001926/2020\|EPI_ISL_452448\|20 | ···················· | ···················· | ···················· | ···················· | NNNNNNNNNNNNNNNNNNNN | NNNNNNNNNNNNNNNNNNNN | 1 |
| C | 5750 hCoV-19/Spain/AN-IBV-001925/2020\|EPI_ISL_452449\|20 | ···················· | ···················· | ···················· | ···················· | NNNNNNNNNNNNNNNNNNNN | NNNNNNNNNNNNNNNNNNNN | 1 |
| C | 5752 hCoV-19/Spain/AN-IBV-001895/2020\|EPI_ISL_452453\|20 | ···················· | ···················· | ···················· | ···················· | NNNNNNNNNNNNNNNNNNNN | NNNNNNNNNNNNNNNNNNNN | 1 |
| C | 5753 hCoV-19/Spain/AN-IBV-001894/2020\|EPI_ISL_452454\|20 | ···················· | ···················· | ···················· | ···················· | NNNNNNNNNNNNNNNNNNNN | NNNNNNNNNNNNNNNNNNNN | 1 |
| C | 5754 hCoV-19/Spain/AN-IBV-001891/2020\|EPI_ISL_452457\|20 | ···················· | ···················· | ···················· | ···················· | NNNNNNNNNNNNNNNNNNNN | N··················· | 1 |
| C | 5755 hCoV-19/Spain/AN-IBV-001889/2020\|EPI_ISL_452459\|20 | ···················· | ···················· | ···················· | ···················· | NNNNNNN············· | ···················· | 1 |
| C | 5756 hCoV-19/Spain/AN-IBV-001888/2020\|EPI_ISL_452460\|20 | ···················· | ···················· | ···················· | ···················· | NNNNNNNNNNNNNNNNNNNN | NNNNNNNNNNNNNNNNNNNN | 1 |
| C | 5757 hCoV-19/Spain/AN-IBV-001887/2020\|EPI_ISL_452461\|20 | ···················· | ···················· | ···················· | ···················· | NNNNNNNNNNNNNNNNNNNN | ···················· | 1 |
| C | 5758 hCoV-19/Spain/AN-IBV-001886/2020\|EPI_ISL_452462\|20 | ···················· | ···················· | ···················· | ···················· | NNNNNNNNNNNNNNNNNNNN | NNNNNNNNNNNNNNN·NNNN | 1 |
| C | 5759 hCoV-19/Spain/AN-IBV-001883/2020\|EPI_ISL_452464\|20 | ···················· | ···················· | ···················· | ···················· | NNNNNNNNNNNNNNNNNNNN | NNNNNNNNNNNNNNN····· | 1 |
| C | 5760 hCoV-19/Spain/AN-IBV-001881/2020\|EPI_ISL_452465\|20 | ···················· | ···················· | ···················· | ···················· | NNNNNNNNNNNNNNNNNNNN | NNNNNNNNNNNNNNNNNNNN | 1 |
| C | 5761 hCoV-19/Spain/AN-IBV-001880/2020\|EPI_ISL_452466\|20 | ···················· | ···················· | ···················· | ···················· | NNNNNNNNNNNNNNNNNNNN | NNNNNNNNNNNNNNNNNNNN | 1 |
| C | 5762 hCoV-19/Spain/AN-IBV-001879/2020\|EPI_ISL_452467\|20 | ···················· | ···················· | ···················· | ···················· | NNNNNNNNNNNNNNNNNNNN | ···················· | 1 |
| C | 5763 hCoV-19/Spain/AN-IBV-001877/2020\|EPI_ISL_452468\|20 | ···················· | ···················· | ···················· | ···················· | NNNNNNNNNNNNNNNNNNNN | NNNNNNNNNNNNNNNNNNNN | 1 |
| C | 5764 hCoV-19/Spain/AN-IBV-001876/2020\|EPI_ISL_452469\|20 | ···················· | ···················· | ···················· | ···················· | NNNNNNNNNNNNNNNNNNNN | NNNNNNNNNNNNNNNNNNNN | 1 |
| C | 5765 hCoV-19/Spain/AN-IBV-001875/2020\|EPI_ISL_452470\|20 | ···················· | ···················· | ···················· | ···················· | NNNNNNNNNNNNNNNNNNNN | NNNNNNNNNNNNNNNNNNNN | 1 |
| C | 5766 hCoV-19/Spain/AN-IBV-001874/2020\|EPI_ISL_452471\|20 | ···················· | ···················· | ···················· | ···················· | NNNNNNNNNNNNNNNNNNNN | NNNNNNNNNNNNNNNNNNNN | 1 |
| C | 5767 hCoV-19/Spain/PV-IBV-001428/2020\|EPI_ISL_452473\|20 | ···················· | ···················· | ···················· | ···················· | NNNNNNNNNNNNNNNNNNNN | NNNNNNNNNNNNNNNNNNNN | 1 |
| C | 5768 hCoV-19/Spain/NC-IBV-001427/2020\|EPI_ISL_452474\|20 | ···················· | ···················· | ···················· | ···················· | NNNNNNNNNNNNNNNNNNNN | NN············N····· | 1 |
| C | 5769 hCoV-19/Spain/NC-IBV-001425/2020\|EPI_ISL_452476\|20 | ···················· | ···················· | ···················· | ···················· | NNNNNNNNNNNNNNNNNNNN | NNNNNNNNNNNNNNNNNNNN | 1 |
| C | 5770 hCoV-19/Spain/NC-IBV-001424/2020\|EPI_ISL_452477\|20 | ···················· | ···················· | ···················· | ···················· | NNNNNNNNNNNNNNNNNNNN | NNNNNNNNNNNNNNNNNNNN | 1 |
| C | 5771 hCoV-19/Spain/NC-IBV-001422/2020\|EPI_ISL_452479\|20 | ···················· | ···················· | ···················· | ···················· | NNNNNNNNNNNNNNNNNNNN | NNNNNNNNNNNNNNNNNNNN | 1 |
| C | 5772 hCoV-19/Spain/PV-IBV-001421/2020\|EPI_ISL_452480\|20 | ···················· | ···················· | ···················· | ···················· | NNNNNNNNNNNNNNNNNNNN | NNNNNNNNNNNNNNNNNNNN | 1 |
| C | 5773 hCoV-19/Spain/NC-IBV-001420/2020\|EPI_ISL_452481\|20 | ···················· | ···················· | ···················· | ···················· | NNNNNNNNNNNNNNNNNNNN | NNNNNNNNNNNNNNNNNNNN | 1 |
| C | 5775 hCoV-19/Spain/NC-IBV-001417/2020\|EPI_ISL_452484\|20 | ···················· | ···················· | ···················· | ···················· | NNNNNNNNNNNNNNNNNNNN | ···················· | 1 |
| C | 5777 hCoV-19/Spain/NC-IBV-001415/2020\|EPI_ISL_452486\|20 | ···················· | ···················· | ···················· | ···················· | NNNNNNNNNNNNNNNNNNNN | ···················· | 1 |
| C | 5778 hCoV-19/Spain/NC-IBV-001414/2020\|EPI_ISL_452487\|20 | ···················· | ···················· | ···················· | ···················· | NNNNNNNNNNNNNNNNNNNN | NNNNNNNNNNNNNNNNNNNN | 1 |
| C | 5779 hCoV-19/Spain/NC-IBV-001413/2020\|EPI_ISL_452488\|20 | ···················· | ···················· | ···················· | ···················· | NNNNNNNNNNNNNNNNNNNN | NNNNNNNNNNNNNNNNNNNN | 1 |
| C | 5780 hCoV-19/Spain/NC-IBV-001411/2020\|EPI_ISL_452490\|20 | ···················· | ···················· | ···················· | ···················· | NNNNNNNNNNNNNNNNNNNN | NNNNNNNNNNNNNNNNNNNN | 1 |
| C | 5781 hCoV-19/Spain/RI-IBV-001410/2020\|EPI_ISL_452491\|20 | ···················· | ···················· | ···················· | ···················· | NNNNNNNNNNNNNNNNNNNN | NNNNNNNNNNNNNNNNNNNN | 1 |
| C | 5782 hCoV-19/Spain/NC-IBV-001409/2020\|EPI_ISL_452492\|20 | ···················· | ···················· | ···················· | ···················· | NNNNNNNNNNNNNNNNNNNN | NNNNNNNNNNNNNNNNNNNN | 1 |
| C | 5783 hCoV-19/Spain/NC-IBV-001408/2020\|EPI_ISL_452493\|20 | ···················· | ···················· | ···················· | ···················· | NNNNNNN············· | ···················· | 1 |
| C | 5784 hCoV-19/Spain/NC-IBV-001406/2020\|EPI_ISL_452495\|20 | ···················· | ···················· | ···················· | ···················· | NNNNNNNNNNNN········ | NNNNNNNNNNNNNNN····· | 1 |
| C | 5785 hCoV-19/Spain/NC-IBV-001405/2020\|EPI_ISL_452496\|20 | ···················· | ·A·················· | ···················· | ···················· | NNNNNNNNNNNNNNNNNNNN | NNNNNNNNNNNNNNNNNNNN | 1 |
| C | 5786 hCoV-19/Spain/NC-IBV-001403/2020\|EPI_ISL_452498\|20 | ···················· | ···················· | ···················· | ···················· | NNNNNNN············· | ···················· | 1 |
| C | 5932 hCoV-19/Spain/AN-IBV-001944/2020\|EPI_ISL_452434\|20 | ···················· | ···················· | ···················· | ···················· | ··NNN··············· | NNNNNNNNNNNNNNN····· | 1 |
| C | 5933 hCoV-19/Spain/AN-IBV-001941/2020\|EPI_ISL_452436\|20 | ···················· | ···················· | ···················· | ···················· | NNNNNNNNNNNNNNNNNNNN | NNNNNNNNNNNNNNNNNNNN | 1 |
| C | 5934 hCoV-19/Spain/AN-IBV-001937/2020\|EPI_ISL_452439\|20 | ···················· | ···················· | ···················· | ···················· | NNNNNNNNNNNNNNNNNNNN | NNNNNNNNNNNNNNNNNNNN | 1 |
| C | 5964 hCoV-19/England/EXET-136185/2020\|EPI_ISL_457205\|20 | ···················· | ···················· | ···················· | ···················· | NNNNNNNNNNNNNNNNNNNN | NNNNNNNNNNNNNNNNNNNN | 1 |
| C | 5967 hCoV-19/England/EXET-136325/2020\|EPI_ISL_457220\|20 | ···················· | ···················· | ···················· | ···················· | NNNNNNNNNNNNNNNNNNNN | NNNNNNNNNNNNNNNNNNNN | 1 |
| C | 5970 hCoV-19/England/EXET-136361/2020\|EPI_ISL_457223\|20 | ···················· | ···················· | ···················· | ···················· | NNNNNNNNNNNNNNNNNNNN | NNNNNNNNNNNNNNNNNNNN | 1 |
| C | 5973 hCoV-19/England/EXET-136686/2020\|EPI_ISL_457229\|20 | ···················· | ···················· | ···················· | ···················· | NNNNNNNNNNNNNNNNNNNN | NNNNNNNNNNNNNNNNNNNN | 1 |
| C | 5974 hCoV-19/England/EXET-13670B/2020\|EPI_ISL_457232\|20 | ···················· | ···················· | ···················· | ···················· | NNNNNNNNNNNNNNNNNNNN | NNNNNNNNNNNNNNNNNNNN | 1 |
| C | 5975 hCoV-19/England/EXET-136765/2020\|EPI_ISL_457233\|20 | ···················· | ···················· | ···················· | ···················· | NNNNNNNNNNNNNNNNNNNN | NNNNNNNNNNNNNNNNNNNN | 1 |
| C | 5976 hCoV-19/England/EXET-136792/2020\|EPI_ISL_457234\|20 | ···················· | ···················· | ···················· | ···················· | NNNNNNNNNNNNNNNNNNNN | NNNNNNNNNNNNNNNNNNNN | 1 |
| C | 5977 hCoV-19/England/EXET-1367FC/2020\|EPI_ISL_457236\|20 | ···················· | ···················· | ···················· | ···················· | NNNNNNNNNNNNNNNNNNNN | NNNNNNNNNNNNNNNNNNNN | 1 |
| C | 5978 hCoV-19/England/EXET-136871/2020\|EPI_ISL_457238\|20 | ···················· | ···················· | ···················· | ···················· | NNNNNNNNNNNNNNNNNNNN | NNNNNNNNNNNNNNNNNNNN | 1 |

| | | | | | | | | | |
|---|---|---|---|---|---|---|---|---|---|
| C | 5982 | hCoV-19/England/EXET-136A11/2020\|EPI_ISL_457243\|20 | ···················· | ···················· | ···················· | ···················· | NNNNNNNNNNNNNNNNNNNN | NNNNNNNNNNNNNNNNNNNN | 1 |
| C | 5986 | hCoV-19/England/EXET-136AE4/2020\|EPI_ISL_457248\|20 | ···················· | ···················· | ···················· | ···················· | NNNNNNNNNNNNNNNNNNNN | NNNNNNNNNNNNNNNNNNNN | 1 |
| C | 6159 | hCoV-19/Northern Ireland/NIRE-FADA8/2020\|EPI_ISL_4 | ···················· | ···················· | ···················· | ···················· | ·NNN················ | NNNNNNNNNNNNNNNNNNNN | 1 |
| C | 6160 | hCoV-19/Northern Ireland/NIRE-FAE1E/2020\|EPI_ISL_4 | ···················· | ···················· | ···················· | ···················· | NNNNN··············· | ···················· | 1 |
| C | 6166 | hCoV-19/Germany/BY-MVP-0098/2020\|EPI_ISL_452104\|20 | ···················· | ···················· | ···················· | ···················· | NNNNNNNNNNNNNNNNNNNN | ············NNN····· | 1 |
| C | 6236 | hCoV-19/India/MH-NIV-11677/2020\|EPI_ISL_452194\|202 | ···················· | ···················· | ···················· | ·········NNNNNN····· | ···················· | ···················· | 1 |
| C | 6255 | hCoV-19/Northern Ireland/NIRE-101E13/2020\|EPI_ISL_ | ···················· | ···················· | ···················· | ···················· | NNNNNNNNNNNNNNNNNNNN | ·············NNN···· | 1 |
| C | 6262 | hCoV-19/Northern Ireland/NIRE-101E9B/2020\|EPI_ISL_ | ···················· | ···················· | ···················· | ···················· | NNNNNNNNNNNNNNNNNNNN | NNNNNNNNNNNNNNNNNNNN | 1 |
| C | 6264 | hCoV-19/Northern Ireland/NIRE-101EB9/2020\|EPI_ISL_ | ···················· | ···················· | ···················· | ···················· | ·NNNNNNNNNNNNNNN···· | NNNNNNNNNNNNNNN····· | 1 |
| C | 6265 | hCoV-19/Northern Ireland/NIRE-101EC8/2020\|EPI_ISL_ | ···················· | ···················· | ···················· | ···················· | NNN······NNNNNNNN··· | ···················· | 1 |
| C | 6271 | hCoV-19/Northern Ireland/NIRE-101F98/2020\|EPI_ISL_ | ···················· | ···················· | ···················· | ···················· | NNNNNNNNNNNNN······· | ···················· | 1 |
| C | 6353 | hCoV-19/England/EXET-1361D0/2020\|EPI_ISL_457209\|20 | ···················· | ···················· | ···················· | ···················· | NNNNNNNNNNNNNNNNNNNN | NNNNNNNNNNNNNNNNNNNN | 1 |
| C | 6354 | hCoV-19/England/EXET-1361EF/2020\|EPI_ISL_457210\|20 | ···················· | ···················· | ···················· | ···················· | NNNNNNNNNNNNNNNNNNNN | NNNNNNNNNNNNNNNNNNNN | 1 |
| C | 6357 | hCoV-19/England/EXET-136264/2020\|EPI_ISL_457213\|20 | ···················· | ···················· | ···················· | ···················· | NNNNNNNNNNNNNNNNNNNN | NNNNNNNNNNNNNNNNNNNN | 1 |
| C | 6375 | hCoV-19/Canada/ON_QGLO-010/2020\|EPI_ISL_459869\|202 | ···················· | ···················· | ···················· | TNNNNN··N··N·NNN···· | ···················· | ···················· | 1 |
| C | 6402 | hCoV-19/Spain/VC-FISABIO-570/2020\|EPI_ISL_447501\|2 | ···················· | ···················· | ···················· | ···················· | ···················· | NNNNNNNNNNNNNNN····· | 1 |
| C | 6415 | hCoV-19/Spain/VC-FISABIO-598/2020\|EPI_ISL_447520\|2 | ···················· | ···················· | ···················· | ···················· | NNNNNNNNNNNNNNNNNNNN | NNNNNNNNNNNNNNNNNNNN | 1 |
| C | 6417 | hCoV-19/Spain/VC-FISABIO-604/2020\|EPI_ISL_447523\|2 | ···················· | ···················· | ···················· | ···················· | ·NNNNNN············· | NNNNNNNNNNNNNNN····· | 1 |
| C | 6622 | hCoV-19/England/NORW-E760D/2020\|EPI_ISL_461805\|202 | ···················· | ···················· | ···················· | ···················· | NNNNNNNNNNNNNNNNNNNN | NNNNNNNNNNNNNNNNNNNN | 1 |
| C | 6623 | hCoV-19/England/NORW-E78DA/2020\|EPI_ISL_461806\|202 | ···················· | ···················· | ···················· | ···················· | NNNNNNNNNNNNNNNNNNNN | NNNNNNNNNNNNNNNNNNNN | 1 |
| C | 6625 | hCoV-19/England/NORW-E8DDA/2020\|EPI_ISL_461810\|202 | ··················T· | ···················· | ···················· | ···················· | NNNNNNNNNNNNNNNNNNNN | NNNNNNNNNNNNNNNNNNNN | 1 |
| C | 6626 | hCoV-19/England/NORW-E8DE9/2020\|EPI_ISL_461811\|202 | ···················· | ···················· | ···················· | ···················· | NNNNN··············· | ···················· | 1 |
| C | 6627 | hCoV-19/England/NORW-E8E40/2020\|EPI_ISL_461812\|202 | ···················· | ···················· | ···················· | ···················· | NNNNNNNNNNNNNNNNNNNN | NNNNNNNNNNNNNNN····· | 1 |
| C | 6628 | hCoV-19/England/NORW-E8E7D/2020\|EPI_ISL_461813\|202 | ···················· | ···················· | ···················· | ···················· | NNNNNNNNNNNNNNNNNNNN | NNNNNNNNNNNNNNNNNNNN | 1 |
| C | 6630 | hCoV-19/England/NORW-E9094/2020\|EPI_ISL_461816\|202 | ···················· | ···················· | ···················· | ···················· | NNNNNNNNNNNNNNNNNNNN | NNNNNNNNNNNNNNNN···· | 1 |
| C | 6631 | hCoV-19/England/NORW-E90A3/2020\|EPI_ISL_461817\|202 | ···················· | ···················· | ···················· | ···················· | NNNNNNNNNNNNN······· | ···················· | 1 |
| C | 6633 | hCoV-19/England/NORW-EA5FE/2020\|EPI_ISL_461857\|202 | ···················· | ···················· | ···················· | ···················· | NNNNNNNNNNNNNNNNNNNN | NNNNNNNNNNNNNNNNNNNN | 1 |
| C | 6634 | hCoV-19/England/NORW-EA619/2020\|EPI_ISL_461858\|202 | ···················· | ···················· | ···················· | ···················· | NNNNNNNNNNNNNNNNNNNN | NNNNNNNNNNNNNNNNNNNN | 1 |
| C | 6635 | hCoV-19/England/NORW-EA646/2020\|EPI_ISL_461860\|202 | ···················· | ···················· | ··········T········· | ···················· | NNNNNNNNNNNNNNNNNNNN | NNNNNNNNNNNNNNNNNNNN | 1 |
| C | 6636 | hCoV-19/England/NORW-EA664/2020\|EPI_ISL_461861\|202 | ···················· | ···················· | ···················· | ···················· | NNNNNNNNNNNNNNNNNNNN | NNNNNNNNNNNNNNNNNNNN | 1 |
| C | 6637 | hCoV-19/England/NORW-EA734/2020\|EPI_ISL_461862\|202 | ···················· | ···················A | ···················· | ···················· | NNNNNNNNNNNNNNNNNNNN | NNNNNNNNNNNNNNNNNNNN | 1 |
| C | 6638 | hCoV-19/England/NORW-EA743/2020\|EPI_ISL_461863\|202 | ···················· | ···················· | ···················· | ···················· | NNNNNNNNNNNNNNNNNNNN | NNNNNNNNNNNNNNNNNNNN | 1 |
| C | 6639 | hCoV-19/England/NORW-EA770/2020\|EPI_ISL_461864\|202 | ···················· | ···················· | ···················· | ···················· | NNNNNNNNNNNNNNNNNNNN | NNNNNNNNNNNNNNNNNNNN | 1 |
| C | 6640 | hCoV-19/England/NORW-EA7BC/2020\|EPI_ISL_461865\|202 | ···················· | ···················· | ···················· | ···················· | NNNNNNNNNNNNNNNNNNNN | NNNNNNNNNNNNNNNNNNNN | 1 |
| C | 6641 | hCoV-19/England/NORW-EA7CB/2020\|EPI_ISL_461866\|202 | ···················· | ···················· | ···················· | ···················· | NNNNNNNNNNNNNNNNNNNN | NNNNNNNNNNNNNNNNNNNN | 1 |
| C | 6642 | hCoV-19/England/NORW-EA7DA/2020\|EPI_ISL_461867\|202 | ···················· | ···················· | ···················· | ···················· | NNNNNNNNNNNNNNNNNNNN | NNNNNNNNNNNNNNNNNNNN | 1 |
| C | 6643 | hCoV-19/England/NORW-EA804/2020\|EPI_ISL_461868\|202 | ···················· | ···················· | ···················· | ···················· | NNNNNNNNNNNNNNNNNNNN | NNNNNNNNNNNNNNNNNNNN | 1 |
| C | 6644 | hCoV-19/England/NORW-EA87D/2020\|EPI_ISL_461870\|202 | ···················· | ···················· | ···················· | ···················· | NNNNNNNNNNNNNNNNNNNN | NNNNNNNNNNNNNNNNNNNN | 1 |
| C | 6645 | hCoV-19/England/NORW-EA8AA/2020\|EPI_ISL_461871\|202 | ···················· | ···················· | ···················· | ···················· | NNNNNNNNNNNNNNNNNNNN | NNNNNNNNNNNNNNNNNNNN | 1 |
| C | 6646 | hCoV-19/England/NORW-EA8D7/2020\|EPI_ISL_461872\|202 | ···················· | ···················· | ···················· | ···················· | NNNNNNNNNNNNNNNNNNNN | NNNNNNNNNNNNNNNNNNNN | 1 |
| C | 6647 | hCoV-19/England/NORW-EA94D/2020\|EPI_ISL_461876\|202 | ···················· | ···················· | ···················· | ···················· | NNNNNNNNNNNNNNNNNNNN | NNNNNNNNNNNNNNNNNNNN | 1 |
| C | 6648 | hCoV-19/England/NORW-EA998/2020\|EPI_ISL_461879\|202 | ···················· | ···················· | ···················· | ···················· | NNNNNNNNNNNNNNNNNNNN | NNNNNNNNNNNNNNNNNNNN | 1 |
| C | 6649 | hCoV-19/England/NORW-EAA0E/2020\|EPI_ISL_461880\|202 | ···················· | ···················· | ···················· | ···················· | NNNNNNNNNNNNNNNNNNNN | NNNNNNNNNNNNNNNNNNNN | 1 |
| C | 6650 | hCoV-19/England/NORW-EAA1D/2020\|EPI_ISL_461881\|202 | ···················· | ···················· | ···················· | ···················· | NNNNNNNNNNNNNNNNNNNN | NNNNNNNNNNNNNNNNNNNN | 1 |
| C | 6651 | hCoV-19/England/NORW-EAA4A/2020\|EPI_ISL_461883\|202 | ···················· | ···················· | ···················· | ···················· | NNNNNNNNNNNNNNNNNNNN | NNNNNNNNNNNNNNNNNNNN | 1 |
| C | 6653 | hCoV-19/England/NORW-EAC44/2020\|EPI_ISL_461885\|202 | ···················· | ···················· | ···················· | ···················· | NNNNNNNNNNNNNNNNNNNN | NNNN················ | 1 |
| C | 6654 | hCoV-19/England/NORW-EB1F9/2020\|EPI_ISL_461887\|202 | ···················· | ···················· | ···················· | ···················· | NNNNNNNNNNNNNNNNNNNN | NNNNNNNNNNNNNNNNNNNN | 1 |
| C | 6655 | hCoV-19/England/NORW-EB205/2020\|EPI_ISL_461888\|202 | ···················· | ···················· | ···················· | ···················· | NNNNNNNNNNNNNNNNNNNN | NNNNNNNNNNNNNNNNNNNN | 1 |
| C | 6656 | hCoV-19/England/NORW-EB223/2020\|EPI_ISL_461890\|202 | ···················· | ···················· | ···················· | ···················· | NNNNNNNNNNNNNNNNNNNN | NNNNNNNNNNNNNNNNNNNN | 1 |
| C | 6658 | hCoV-19/Spain/VC-FISABIO-610/2020\|EPI_ISL_447528\|2 | ···················· | ···················· | ···················· | ···················· | NNNNNNNNNNNNNNNNNNNN | NNNNNNNNNNNNNNNNNNNN | 1 |
| C | 6660 | hCoV-19/Spain/VC-FISABIO-612/2020\|EPI_ISL_447530\|2 | ···················· | ···················· | ···················· | ···················· | NNNNNNNNNNNNNNNNNNNN | NNNNNNNNNNNNNNNNNNNN | 1 |
| C | 6696 | hCoV-19/India/MH-NIV-6311/2020\|EPI_ISL_452206\|2020 | ···················· | ···················· | ···················· | NNNNNNNNNNNNNNNT···· | ···················· | ···················· | 1 |
| C | 6728 | hCoV-19/England/20109006304/2020\|EPI_ISL_452256\|20 | ···················· | ···················· | ···················· | ···················· | NNNNNNNNNNNNNNNNNNNN | ···················· | 1 |
| C | 6773 | hCoV-19/England/NORW-E8EB9/2020\|EPI_ISL_453561\|202 | ···················· | ···················· | ···················· | ···················· | ···················· | NNNNN··············· | 1 |
| C | 6903 | hCoV-19/England/NORW-E910A/2020\|EPI_ISL_461821\|202 | ···················· | ···················· | ···················· | ···················· | NNNNNNNNNNNNNNNNNNNN | NNNN················ | 1 |
| C | 6909 | hCoV-19/England/NORW-E939B/2020\|EPI_ISL_461829\|202 | ···················· | ···················· | ···················· | ···················· | NNNNNNNNNNNNNNNNNNNN | NNNNNNNNNNNNNNNNNNNN | 1 |
| C | 6910 | hCoV-19/England/NORW-E98E7/2020\|EPI_ISL_461836\|202 | ···················· | ···················· | ···················· | ···················· | NNNN················ | ···················· | 1 |
| C | 6912 | hCoV-19/England/NORW-E9AB4/2020\|EPI_ISL_461844\|202 | ···················· | ···················· | ···················· | ···················· | NNNNNNNNNNNNNNNNNNNN | ···················· | 1 |
| C | 6914 | hCoV-19/Northern Ireland/NIRE-FAFFD/2020\|EPI_ISL_4 | ···················· | ···················· | ···················· | ···················· | NNNN················ | ···················· | 1 |
| C | 6915 | hCoV-19/Northern Ireland/NIRE-FB16F/2020\|EPI_ISL_4 | ···················· | ···················· | ···················· | ···················· | NNNNNNNNNNNNNNNNNNNN | NNNNNNNNNNNNNNNNNNNN | 1 |
| C | 6916 | hCoV-19/Northern Ireland/NIRE-FB19C/2020\|EPI_ISL_4 | ···················· | ···················· | ···················· | ···················· | NNNNNNNNNNNNNNNNNNNN | NNNNNNNNNNNNNNNNNNNN | 1 |
| C | 6917 | hCoV-19/Northern Ireland/NIRE-FB27B/2020\|EPI_ISL_4 | ···················· | ···················· | ···················· | ···················· | NNNNNNNNNNNNNNNNNNNN | NNNNNNNNNNNNNNNNNNNN | 1 |
| C | 6918 | hCoV-19/Northern Ireland/NIRE-FB615/2020\|EPI_ISL_4 | ···················· | ···················· | ···················· | ···················· | NNNNNNNNNNNNNNNNNNNN | NNNNNNNNNNNNNNNNNNNN | 1 |
| C | 6919 | hCoV-19/Northern Ireland/NIRE-FB633/2020\|EPI_ISL_4 | ···················· | ···················· | ···················· | ···················· | NNNNNNNNNNNNNNNNNNNN | NNNNNNNNNNNNNNNNNNNN | 1 |
| C | 6920 | hCoV-19/Northern Ireland/NIRE-FB67F/2020\|EPI_ISL_4 | ···················· | ···················· | ···················· | ···················· | ·NNNNNNNNNNNNNNNNNNN | NNNNNNNNNNNNNNNNNNNN | 1 |
| C | 6921 | hCoV-19/Northern Ireland/NIRE-FB68E/2020\|EPI_ISL_4 | ···················· | ···················· | ···················· | ···················· | NNNNNNNNNNNNNNNNNNNN | NNNNNNNNNNNNNNNNNNNN | 1 |
| C | 6922 | hCoV-19/Northern Ireland/NIRE-FB6CA/2020\|EPI_ISL_4 | ···················· | ···················· | ···················· | ···················· | NNNNNNNNNNNNNNNNNNNN | NNNNNNNNNNNNNNNNNNNN | 1 |
| C | 6923 | hCoV-19/Northern Ireland/NIRE-FB6D9/2020\|EPI_ISL_4 | ···················· | ···················· | ···················· | ···················· | NNNNNNNNNNNNNNNNNNNN | NNNNNNNNNNNNNNNN···· | 1 |
| C | 6924 | hCoV-19/Northern Ireland/NIRE-FB6E8/2020\|EPI_ISL_4 | ···················· | ···················· | ···················· | ···················· | NNNNNNNNNNNNNNNNNNNN | NNNNNNNNNNNNNNNNNNNN | 1 |
| C | 6925 | hCoV-19/Northern Ireland/NIRE-FB6F7/2020\|EPI_ISL_4 | ···················· | ···················· | ···················· | ···················· | NNNNNNNNNNNNNNNNNNNN | NNNNNNNNNNNNNNNNNNNN | 1 |
| C | 6926 | hCoV-19/Northern Ireland/NIRE-FB712/2020\|EPI_ISL_4 | ···················· | ···················· | ···················· | ···················· | ···················· | NNNNNNNNNNNNNNNN···· | 1 |
| C | 6929 | hCoV-19/Northern Ireland/NIRE-FB74F/2020\|EPI_ISL_4 | ···················· | ···················· | ···················· | ···················· | NNNNNNNNNNNNNNNNNNNN | NNNNNNNNNNNNNNNNNNNN | 1 |
| C | 6930 | hCoV-19/Northern Ireland/NIRE-FB75E/2020\|EPI_ISL_4 | ···················· | ···················· | ···················· | ···················· | NNNN················ | ···················· | 1 |
| C | 6931 | hCoV-19/Northern Ireland/NIRE-FB77C/2020\|EPI_ISL_4 | ···················· | ···················· | ···················· | ···················· | NNNNNNNNNNNNNNNNNNNN | NNNNNNNNNNNNNNNNNNNN | 1 |
| C | 6932 | hCoV-19/Northern Ireland/NIRE-FB78B/2020\|EPI_ISL_4 | ···················· | ···················· | ···················· | ···················· | NNNNNNNNNNNNNNNNNNNN | NNNNNNNNNNNNNNNNNNNN | 1 |
| C | 6957 | hCoV-19/Scotland/EDB5376/2020\|EPI_ISL_457012\|2020- | ···················· | ···················· | ···················· | ···················· | NNNNNNNNNNNNNNNNNNNN | NNNNNNNNNNNNNNNNNNNN | 1 |
| C | 7000 | hCoV-19/England/EXET-13558A/2020\|EPI_ISL_457071\|20 | ···················· | ···················· | ···················· | ···················· | NNNNNNNNNNNNNNNNNNNN | NNNNNNNNNNNNNNNNNNNN | 1 |
| C | 7181 | hCoV-19/Northern Ireland/NIRE-FB1BA/2020\|EPI_ISL_4 | ···················· | ···················· | ···················· | ···················· | ·NNNNNNNNNNNNNNNNNNN | ···················· | 1 |
| C | 7183 | hCoV-19/Northern Ireland/NIRE-FB211/2020\|EPI_ISL_4 | ···················· | ···················· | ···················· | ···················· | NNNNNNNNNNNNNNNNNNNN | NNNNNNNNNNNNNNNNNNNN | 1 |
| C | 7185 | hCoV-19/Northern Ireland/NIRE-FB26C/2020\|EPI_ISL_4 | ···················· | ···················· | ···················· | ···················· | NNNNNNNNNNNNNNNNNNNN | ···················· | 1 |

| | | | | | | | | | |
|---|---|---|---|---|---|---|---|---|---|
| C | 7199 | hCoV-19/England/NORW-E6F9A/2020\|EPI_ISL_448994\|202 | ···················· | ···················· | ···················· | ···················· | NNNN················ | ···················· | 1 |
| C | 7202 | hCoV-19/England/NORW-E703C/2020\|EPI_ISL_448998\|202 | ···················· | ···················· | ···················· | ···················· | NNNNNNNNNNNNNNNNNNNN | ···················· | 1 |
| C | 7239 | hCoV-19/Scotland/EDB5313/2020\|EPI_ISL_457000\|2020- | ···················· | ···················· | ···················· | ···················· | NNNNNNNNNNNNNNNNNNNN | NNNNNNNNNNNNNNNNNNNN | 1 |
| C | 7479 | hCoV-19/England/NORW-E86B1/2020\|EPI_ISL_449175\|202 | ···················· | ···················· | ···················· | ···················· | NNNN················ | ···················· | 1 |
| C | 7620 | hCoV-19/Spain/PV-IBV-000715/2020\|EPI_ISL_452753\|20 | ···················· | ···················· | ···················· | ···················· | NNNNNNNNNNNNNNNNNNNN | NNNNNNNNNNNNNNNNNNNN | 1 |
| C | 7621 | hCoV-19/Spain/PV-IBV-000714/2020\|EPI_ISL_452754\|20 | ···················· | ···················· | ···················· | ···················· | ·NNNNNN············· | ···················· | 1 |
| C | 7622 | hCoV-19/Spain/PV-IBV-000712/2020\|EPI_ISL_452755\|20 | ···················· | ···················· | ···················· | ···················· | NNNNNNN············· | ···················· | 1 |
| C | 7623 | hCoV-19/Spain/PV-IBV-000707/2020\|EPI_ISL_452759\|20 | ···················· | ···················· | ···················· | ···················· | ··NNNNN············· | ···················· | 1 |
| C | 7624 | hCoV-19/Spain/PV-IBV-000705/2020\|EPI_ISL_452761\|20 | ···················· | ···················· | ···················· | ···················· | NNNNNNNNNNNNNNNNNNNN | NNNNNNNNNNNNNNNNNNNN | 1 |
| C | 7625 | hCoV-19/Spain/PV-IBV-000704/2020\|EPI_ISL_452762\|20 | ···················· | ···················· | ···················· | ···················· | NNNNNNNNNNNNNNNNNNNN | NNNNNNNNNNNNNNNNN··· | 1 |
| C | 7627 | hCoV-19/Spain/PV-IBV-000699/2020\|EPI_ISL_452765\|20 | ···················· | ···················· | ···················· | ···················· | NNNNNNNNNNNNNNNNNNNN | NNNNNNNNNNNNNNNNNNNN | 1 |
| C | 7628 | hCoV-19/Spain/PV-IBV-000698/2020\|EPI_ISL_452766\|20 | ···················· | ···················· | ···················· | ···················· | NNNNNNNNNNNNNNNNNNNN | NNNNNNNNNNNNNNNNNNNN | 1 |
| C | 7629 | hCoV-19/Spain/PV-IBV-000697/2020\|EPI_ISL_452767\|20 | ···················· | ···················· | ···················· | ···················· | NNNNNNNNNNNNNNNNNNNN | NNNNNNNNNNNNNNN····· | 1 |
| C | 7630 | hCoV-19/Spain/PV-IBV-000696/2020\|EPI_ISL_452768\|20 | ···················· | ···················· | ···················· | ···················· | NNNNNNNNNNNNNNNNNNNN | NNNNNNNNNNNNNNNNNNNN | 1 |
| C | 7631 | hCoV-19/Spain/PV-IBV-000695/2020\|EPI_ISL_452769\|20 | ···················· | ···················· | ···················· | ···················· | ·NNNNNNNNNNNNNNNNNNN | NNNNNNN············· | 1 |
| C | 7632 | hCoV-19/Spain/PV-IBV-000694/2020\|EPI_ISL_452770\|20 | ···················· | ···················· | ···················· | ···················· | NNNNNNNNNNNNNNNNNNNN | NNNNNNNNNNNNNNNNNNNN | 1 |
| C | 7633 | hCoV-19/Spain/PV-IBV-000693/2020\|EPI_ISL_452771\|20 | ···················· | ···················· | ···················· | ···················· | ·NNNNNNNNNNN·NNNN··· | ···················· | 1 |
| C | 7636 | hCoV-19/Spain/PV-IBV-000690/2020\|EPI_ISL_452774\|20 | ···················· | ···················· | ···················· | ···················· | NNNNNNNNNNNNNNNNNNNN | NNNNNNNNNNNNNNN··NNN | 1 |
| C | 7637 | hCoV-19/Spain/PV-IBV-000689/2020\|EPI_ISL_452775\|20 | ···················· | ···················· | ···················· | ···················· | NNNNNNNNNNNNNNNNNNNN | NNNNNNNNNNNNNNNNNNNN | 1 |
| C | 7639 | hCoV-19/Spain/PV-IBV-000678/2020\|EPI_ISL_452780\|20 | ···················· | ···················· | ···················· | ···················· | NNNNNNN············· | ···················· | 1 |
| C | 7640 | hCoV-19/Spain/PV-IBV-000670/2020\|EPI_ISL_452782\|20 | ···················· | ···················· | ···················· | ···················· | NNNNNNNNNNNNNNNNNNNN | NNNNNNNNNNNNNNNNNNNN | 1 |
| C | 7641 | hCoV-19/Spain/PV-IBV-000669/2020\|EPI_ISL_452783\|20 | ···················· | ···················· | ···················· | ···················· | NNNNNNNNNNNNNNNNNNNN | NNNNNNNNNNNNNNNNNNNN | 1 |
| C | 7642 | hCoV-19/Spain/PV-IBV-000666/2020\|EPI_ISL_452784\|20 | ···················· | ···················· | ···················· | ···················· | NNNNNNNNNNNNNNNNNNNN | NNNNNNNNNNNNNNNNNNNN | 1 |
| C | 7643 | hCoV-19/Spain/PV-IBV-000665/2020\|EPI_ISL_452785\|20 | ···················· | ···················· | ···················· | ···················· | ·NNNNNNNNNNNNNNNNNNN | NNNNNNNNNNNNNNNNNNNN | 1 |
| C | 7644 | hCoV-19/Spain/PV-IBV-000664/2020\|EPI_ISL_452786\|20 | ···················· | ···················· | ···················· | ···················· | ·NNNNNNNNNNNNNNNNNNN | NNNNNNNNNNNNNNNNNNNN | 1 |
| C | 7723 | hCoV-19/Spain/PV-IBV-000851/2020\|EPI_ISL_452700\|20 | ···················· | ···················· | ···················· | ···················· | NNNNNNNNNNNNNNNNNNNN | NN·················· | 1 |
| C | 7726 | hCoV-19/Spain/PV-IBV-000842/2020\|EPI_ISL_452704\|20 | ···················· | ···················· | ···················· | ···················· | NNNNNNNNNNNNNNNNNNNN | NNNNNNNNNNNNNNNNNNNN | 1 |
| C | 7727 | hCoV-19/Spain/PV-IBV-000835/2020\|EPI_ISL_452707\|20 | ···················· | ···················· | ···················· | ···················· | NNNNNNNNNNNNNNNNNNNN | NNNNNNNNNNNNNNNNNNNN | 1 |
| C | 7728 | hCoV-19/Spain/PV-IBV-000830/2020\|EPI_ISL_452708\|20 | ···················· | ···················· | ···················· | ···················· | ·NNNNNN············· | ···················· | 1 |
| C | 7729 | hCoV-19/Spain/PV-IBV-000829/2020\|EPI_ISL_452709\|20 | ···················· | ···················· | ···················· | ···················· | ···················· | NNNNNNNNNNNNNNN····· | 1 |
| C | 7730 | hCoV-19/Spain/PV-IBV-000822/2020\|EPI_ISL_452710\|20 | ···················· | ···················· | ···················· | ···················· | NNNNNNNNNNNNNNNNNNNN | NNNNNNNNNNNNNNNNNNNN | 1 |
| C | 7731 | hCoV-19/Spain/PV-IBV-000817/2020\|EPI_ISL_452711\|20 | ···················· | ···················· | ···················· | ···················· | NNNNNNNNNNNNNNNNNNNN | NNNNNNNNNNNNNNNNNNNN | 1 |
| C | 7732 | hCoV-19/Spain/PV-IBV-000816/2020\|EPI_ISL_452712\|20 | ···················· | ···················· | ···················· | ···················· | NNNNNNNNNNNNNNNNNNNN | NNNNNNNNNNNNNNNNNNNN | 1 |
| C | 7733 | hCoV-19/Spain/PV-IBV-000815/2020\|EPI_ISL_452713\|20 | ···················· | ···················· | ···················· | ···················· | NNNNNNNNNNNNNNNNNNNN | NNNNNNNNNNNNNNNNNNNN | 1 |
| C | 7734 | hCoV-19/Spain/PV-IBV-000814/2020\|EPI_ISL_452714\|20 | ···················· | ···················· | ···················· | ···················· | NNNNNNNNNNNNNNNNNNNN | ···················· | 1 |
| C | 7736 | hCoV-19/Spain/PV-IBV-000806/2020\|EPI_ISL_452716\|20 | ···················· | ···················· | ···················· | ···················· | NNNNNNNNNNNNNNNNNNNN | NNNNNNNNNNNNNNNNNNNN | 1 |
| C | 7739 | hCoV-19/Spain/PV-IBV-000796/2020\|EPI_ISL_452719\|20 | ···················· | ···················· | ···················· | ···················· | NNNNNNNNNNNNNNNNNNNN | NNNNNNNNNNNNNNNNNNNN | 1 |
| C | 7740 | hCoV-19/Spain/PV-IBV-000794/2020\|EPI_ISL_452720\|20 | ···················· | ···················· | ···················· | ···················· | NNNNNNNNNNNNNNNNNNNN | NNNNNNNNNNNNNNNNNNNN | 1 |
| C | 7743 | hCoV-19/Spain/PV-IBV-000786/2020\|EPI_ISL_452723\|20 | ···················· | ···················· | ···················· | ···················· | NNNNNNNNNNNN········ | NNNNNNNNNNNNNNN····· | 1 |
| C | 7744 | hCoV-19/Spain/PV-IBV-000784/2020\|EPI_ISL_452724\|20 | ···················· | ···················· | ···················· | ···················· | ···················· | NNNNNNNNNNNNNNN····· | 1 |
| C | 7745 | hCoV-19/Spain/PV-IBV-000783/2020\|EPI_ISL_452725\|20 | ···················· | ···················· | ···················· | ···················· | NNNNNNNNNNNNNNNNNNNN | NNNNNNNNNNNNNNNNNNNN | 1 |
| C | 7747 | hCoV-19/Spain/PV-IBV-000778/2020\|EPI_ISL_452727\|20 | ···················· | ···················· | ···················· | ···················· | NNNNNNNNNNNNNNNNNNNN | NNNNNNNNNNNNNNNNNNNN | 1 |
| C | 7748 | hCoV-19/Spain/PV-IBV-000775/2020\|EPI_ISL_452728\|20 | ···················· | ···················· | ···················· | ···················· | NNNNNNNNNNNNNNNNNNNN | NNNNNNNNNNNNNNNNNNNN | 1 |
| C | 7749 | hCoV-19/Spain/PV-IBV-000769/2020\|EPI_ISL_452729\|20 | ···················· | ···················· | ···················· | ···················· | NNNNNNNNNNNNNNNNNNNN | NNNNNNNNNNNNNNNNNNNN | 1 |
| C | 7750 | hCoV-19/Spain/PV-IBV-000766/2020\|EPI_ISL_452730\|20 | ···················· | ···················· | ···················· | ···················· | NNNNNNNNNNNNNNNNNNNN | ···················· | 1 |
| C | 7751 | hCoV-19/Spain/PV-IBV-000760/2020\|EPI_ISL_452731\|20 | ···················· | ···················· | ···················· | ···················· | ·NNNNNNNNNNN········ | N··················· | 1 |
| C | 7752 | hCoV-19/Spain/PV-IBV-000759/2020\|EPI_ISL_452732\|20 | ···················· | ···················· | ···················· | ···················· | NNNNNNNNNNNNNNNNNNNN | NNNNNNNNNNNNNNNNNNNN | 1 |
| C | 7753 | hCoV-19/Spain/PV-IBV-000758/2020\|EPI_ISL_452733\|20 | ···················· | ···················· | ···················· | ···················· | NNNNNNNNNNNNNNNNNNNN | NNNNNNNNNNNNNNNNNNNN | 1 |
| C | 7755 | hCoV-19/Spain/PV-IBV-000752/2020\|EPI_ISL_452736\|20 | ···················· | ···················· | ···················· | ···················· | NNNNNNNNNNNNNNNNNNNN | NNNNNNNNNNNNNNNNNNNN | 1 |
| C | 7756 | hCoV-19/Spain/PV-IBV-000751/2020\|EPI_ISL_452737\|20 | ···················· | ···················· | ···················· | ···················· | NNNNNNNNNNNNNNNNNNNN | NNNNNNNNNNNNNNNNNNNN | 1 |
| C | 7758 | hCoV-19/Spain/PV-IBV-000749/2020\|EPI_ISL_452739\|20 | ···················· | ···················· | ···················· | ···················· | NNNNNNNNNNNNNNNNNNNN | NNNNNNNNNNNNNNN····· | 1 |
| C | 7759 | hCoV-19/Spain/PV-IBV-000748/2020\|EPI_ISL_452740\|20 | ···················· | ···················· | ···················· | ···················· | ··NNNNN············· | ···················· | 1 |
| C | 7760 | hCoV-19/Spain/PV-IBV-000744/2020\|EPI_ISL_452741\|20 | ···················· | ···················· | ···················· | ···················· | NNNNNNNNNNNNNNNNNNNN | NNNNNNNNNNNNNNN····· | 1 |
| C | 7762 | hCoV-19/Spain/PV-IBV-000737/2020\|EPI_ISL_452743\|20 | ···················· | ···················· | ···················· | ···················· | NNNNNNN·NNNNN·····NN | ···················· | 1 |
| C | 7764 | hCoV-19/Spain/PV-IBV-000730/2020\|EPI_ISL_452745\|20 | ···················· | ···················· | ···················· | ···················· | ···················· | NNNNNNNNNNNNNNN····· | 1 |
| C | 7765 | hCoV-19/Spain/PV-IBV-000729/2020\|EPI_ISL_452746\|20 | ···················· | ···················· | ···················· | ···················· | ···················· | NNNNN·······NNN····· | 1 |
| C | 7769 | hCoV-19/Spain/PV-IBV-000718/2020\|EPI_ISL_452750\|20 | ···················· | ···················· | ···················· | ···················· | NNNNNNNNNNNNNNNNNNNN | ······NNNNNNNNN····· | 1 |
| C | 7806 | hCoV-19/New Zealand/20VR1781/2020\|EPI_ISL_456250\|2 | ···················· | ···················· | ···················· | ···················· | NNNNNNNNNNNNNNNNNNNN | NNNNNNNNNNNNNNNNNNNN | 1 |
| C | 7845 | hCoV-19/Australia/VIC1360/2020\|EPI_ISL_456427\|2020 | ···················· | ···················· | ···················· | ···················· | NNNNNNNNNNNNNNNNNNNN | NNNNNNNNNNNNNNNNNNNN | 1 |
| C | 7915 | hCoV-19/USA/MD-HP00094/2020\|EPI_ISL_457759\|2020-04 | ···················· | ···················· | ···················· | ···················· | NNNNNNNNNNNNNNNNNNNN | NNNNNNNNNNNNNNNNNNNN | 1 |
| C | 7965 | hCoV-19/Scotland/EDB3956/2020\|EPI_ISL_449260\|2020- | ···················· | ···················· | ···················· | ···················· | NNNNNNNNNNNNNNNNNNNN | NNNNNNNNNNNNNNNNNNNN | 1 |
| C | 7966 | hCoV-19/Scotland/EDB3961/2020\|EPI_ISL_449261\|2020- | ···················· | ···················· | ···················· | ···················· | ··N········N··N····· | ···N···NN····NNN··NN | 1 |
| C | 7967 | hCoV-19/Scotland/EDB3972/2020\|EPI_ISL_449264\|2020- | ···················· | ···················· | ···················· | ···················· | NNNNNNNNNNNNNNNNNNNN | NNNNNNNNNNNNNNNNNNNN | 1 |
| C | 7968 | hCoV-19/Scotland/EDB3973/2020\|EPI_ISL_449265\|2020- | ···················· | ···················· | ···················· | ···················· | ··NN·NNN·NN·NNNNNNNN | NNNN··NNN··NNNN····· | 1 |
| C | 7969 | hCoV-19/Scotland/EDB3983/2020\|EPI_ISL_449271\|2020- | ···················· | ·A·················· | ···················· | ···················· | NNNNNNNNNNNNNNNNNNNN | NNNNNNNNNNNNNNNNNNNN | 1 |
| C | 7970 | hCoV-19/Scotland/EDB3986/2020\|EPI_ISL_449272\|2020- | ···················· | ·A·················· | ···················· | ···················· | NNNNNNNNNNNNNNNNNNNN | NNNNNNNNNNNNNNNNNNNN | 1 |
| C | 7971 | hCoV-19/Scotland/EDB4098/2020\|EPI_ISL_449274\|2020- | ···················· | ···················A | ···················· | ···················· | NNNNNNNNNNNNNNNNNNNN | NNNNNNNNNNNNNNNNNNNN | 1 |
| C | 7976 | hCoV-19/Scotland/EDB4175/2020\|EPI_ISL_449279\|2020- | ···················· | ···················A | ···················· | ···················· | NNNNNNNNNNNNNNNNNNNN | NNNNNNNNNNNNNNNNNNNN | 1 |
| C | 7978 | hCoV-19/Scotland/EDB4202/2020\|EPI_ISL_449281\|2020- | ···················· | ···················A | ···················· | ···················· | NNNNNNNNNNNNNNNNNNNN | NNNNNNNNNNNNNNNNNNNN | 1 |
| C | 7980 | hCoV-19/Scotland/EDB4216/2020\|EPI_ISL_449283\|2020- | ···················· | ···················A | ···················· | ···················· | NNNNNNNNNNNNNNNNNNNN | NNNNNNNNNNNNNNNNNNNN | 1 |
| C | 7984 | hCoV-19/Scotland/EDB4279/2020\|EPI_ISL_449289\|2020- | ···················· | ···················A | ···················· | ···················· | NNNNNNNNNNNNNNNNNNNN | NNNNNNNNNNNNNNNNNNNN | 1 |
| C | 7987 | hCoV-19/Scotland/EDB4310/2020\|EPI_ISL_449292\|2020- | ···················· | ···················A | ···················· | ···················· | NNNNNNNNNNNNNNNNNNNN | NNNNNNNNNNNNNNNNNNNN | 1 |
| C | 7988 | hCoV-19/Scotland/EDB4318/2020\|EPI_ISL_449293\|2020- | ···················· | ···················A | ···················· | ···················· | ··N···········NNNN·· | NN····N············· | 1 |
| C | 7990 | hCoV-19/Scotland/EDB4338/2020\|EPI_ISL_449295\|2020- | ···················· | ···················A | ···················· | ···················· | NNNNNNNNNNNNNNNNNNNN | NNNNNNNNNNNNNNNNNNNN | 1 |
| C | 7991 | hCoV-19/Scotland/EDB4361/2020\|EPI_ISL_449296\|2020- | ···················· | ···················A | ···················· | ···················· | ·····NNN····N·NN···· | NN·N··NNNN···NN····· | 1 |
| C | 8145 | hCoV-19/England/CAMB-1B05B4/2020\|EPI_ISL_452896\|20 | ···················· | ···················· | ···················· | ···················· | NNNNNNNNNNNNNNNNNNNN | NNNNNNNNNNNNNNNNNNNN | 1 |
| C | 8155 | hCoV-19/Australia/VIC1359/2020\|EPI_ISL_456410\|2020 | ···················· | ···················· | ···················· | ···················· | NNNNNNNNNNNNNNNNNNNN | NNNNNNNNNNNNNNNNNNNN | 1 |
| C | 8156 | hCoV-19/Australia/VIC1365/2020\|EPI_ISL_456411\|2020 | ···················· | ···················· | ···················· | ···················· | NN·················· | NNNN···NNNNNNNN····· | 1 |
| C | 8157 | hCoV-19/Australia/VIC1372/2020\|EPI_ISL_456414\|2020 | ···················· | ···················· | ···················· | ···················· | NNNNNNNNNNNNNNNNNNNN | NNNNNNNNNNNNNNNNNNNN | 1 |
| C | 8158 | hCoV-19/Australia/VIC1378/2020\|EPI_ISL_456417\|2020 | ···················· | ···················· | ···················· | ···················· | NNNNNNNNNNNNNNNNNNNN | NNNNNNNNNNNNNNNNNNNN | 1 |

| | | | | | | | | |
|---|---|---|---|---|---|---|---|---|
| C | 8159 hCoV-19/Australia/VIC1382/2020\|EPI_ISL_456419\|2020 | ···················· | ···················· | ···················· | ···················· | NNNNNNNNNNNNNNNNNNNN | NNNNNNNNNNNNNNNNNNNN | 1 |
| C | 8160 hCoV-19/Australia/VIC1364/2020\|EPI_ISL_456421\|2020 | ···················· | ···················· | ···················· | ···················· | NNNNNNN············· | NNNN·M·············· | 1 |
| C | 8161 hCoV-19/Australia/VIC1408/2020\|EPI_ISL_456426\|2020 | ···················· | ···················· | ···················· | ···················· | NNNNNNN············· | ···················· | 1 |
| C | 8162 hCoV-19/Australia/VIC1410/2020\|EPI_ISL_456429\|2020 | ···················· | ···················· | ···················· | ···················· | NNNNNNNNNNNNNNNNNNNN | NNNNNNNNNNNNNNNNNNNN | 1 |
| C | 8163 hCoV-19/Australia/VIC1400/2020\|EPI_ISL_456433\|2020 | ···················· | ···················· | ···················· | ···················· | NNNNNNNNNNNNNNNNNNNN | NNNNNNNNNNNNNNNNNNNN | 1 |
| C | 8164 hCoV-19/Australia/VIC1428/2020\|EPI_ISL_456435\|2020 | ···················· | ···················· | ···················· | ···················· | ·NNNNNNNNNNNNNNNNNNN | ···················· | 1 |
| C | 8184 hCoV-19/USA/MD-HP00077/2020\|EPI_ISL_457755\|2020-03 | ···················· | ···················· | ···················· | ···················· | NNNNNNNNNNNNNNNNNNNN | NNNNNNNNNNNNNNNNNNNN | 1 |
| C | 8185 hCoV-19/USA/MD-HP00086/2020\|EPI_ISL_457756\|2020-03 | ···················· | ···················· | ···················· | ···················· | NNNNNNNNNNNNNNNNNNNN | NNNNNNNNNNNNNNNNNNNN | 1 |
| C | 8186 hCoV-19/USA/DC-HP00091/2020\|EPI_ISL_457757\|2020-03 | ···················· | ···················· | ···················· | ···················· | NNNNNNNNNNNNNNNNNNNN | NNNNNNNNNNNNNNNNNNNN | 1 |
| C | 8187 hCoV-19/USA/MD-HP00093/2020\|EPI_ISL_457758\|2020-03 | ···················· | ···················· | ···················· | ···················· | NNNNNNNNNNNNNNNNNNNN | NNNNNNNNNNNNNNNNNNNN | 1 |
| C | 8189 hCoV-19/USA/MD-HP00097/2020\|EPI_ISL_457761\|2020-03 | ···················· | ···················· | ···················· | ···················· | NNNNNNNNNNNNNNNNNNNN | NNNNNNNNNNNNNNNNNNNN | 1 |
| C | 8190 hCoV-19/USA/MD-HP00106/2020\|EPI_ISL_457764\|2020-03 | ···················· | ···················· | ···················· | ···················· | NNNNNNNNNNNNNNNNNNNN | NNNNNNNNNNNNNNNNNNNN | 1 |
| C | 8191 hCoV-19/USA/MD-HP00107/2020\|EPI_ISL_457765\|2020-03 | ···················· | ···················· | ···················· | ···················· | NNNNNNNNNNNNNNNNNNNN | NNNNNNNNNNNNNNNNNNNN | 1 |
| C | 8312 hCoV-19/England/CAMB-1AEDA2/2020\|EPI_ISL_452862\|20 | ···················· | ···················· | ···················· | ···················· | NNNNNNNNNNNNNNNNNNNN | NNNNNNNNNNNNNNNNNNNN | 1 |
| C | 8532 hCoV-19/Spain/NC-IBV-001391/2020\|EPI_ISL_452510\|20 | ···················· | ···················· | ···················· | ···················· | NNNNNNNNNNNNNNNNNNNN | NNNNNNNNNNNNNNN····· | 1 |
| C | 8533 hCoV-19/Spain/NC-IBV-001388/2020\|EPI_ISL_452513\|20 | ···················· | ···················· | ···················· | ···················· | NNNNNNNNNNNNNNNNNNNN | NNNNNNNNNNNNNNNNNNNN | 1 |
| C | 8534 hCoV-19/Spain/NC-IBV-001387/2020\|EPI_ISL_452514\|20 | ···················· | ···················· | ···················· | ···················· | NNNNNNNNNNNNNNNNNNNN | ···················· | 1 |
| C | 8536 hCoV-19/Spain/NC-IBV-001384/2020\|EPI_ISL_452517\|20 | ···················· | ···················· | ···················· | ···················· | NNNNNNNNNNNNNNNNNNNN | NNNNNNNNNNNNNNNNNNNN | 1 |
| C | 8537 hCoV-19/Spain/NC-IBV-001383/2020\|EPI_ISL_452518\|20 | ···················· | ·············A······ | ···················· | ···················· | NNNNNNN············· | ···················· | 1 |
| C | 8538 hCoV-19/Spain/NC-IBV-001382/2020\|EPI_ISL_452519\|20 | ···················· | ···················· | ···················· | ···················· | NNNNNNNNNNNNNNNNNNNN | ··············N····· | 1 |
| C | 8540 hCoV-19/Spain/NC-IBV-001380/2020\|EPI_ISL_452521\|20 | ···················· | ···················· | ···················· | ···················· | NNNNNNNNNNNNNNNNNNNN | NNNNNNNNNNNNNNNNNNNN | 1 |
| C | 8542 hCoV-19/Spain/NC-IBV-001377/2020\|EPI_ISL_452524\|20 | ···················· | ···················· | ···················· | ···················· | NNNNNNNNNNNNNNNNNNNN | NNNNNNNNNNNNNNNNNNNN | 1 |
| C | 8543 hCoV-19/Spain/RI-IBV-001375/2020\|EPI_ISL_452526\|20 | ···················· | ···················· | ···················· | ···················· | NNNNNNNNNNNNNNNNNNNN | NNNNNNNNNNNNNNNNNNNN | 1 |
| C | 8544 hCoV-19/Spain/NC-IBV-001374/2020\|EPI_ISL_452527\|20 | ···················· | ···················· | ···················· | ···················· | NNNNNNNNNNNNNNNNNNNN | ···················· | 1 |
| C | 8545 hCoV-19/Spain/NC-IBV-001373/2020\|EPI_ISL_452528\|20 | ···················· | ···················· | ···················· | ···················· | NNNNNNNNNNNNNNNNNNNN | NNNNNNNNNNNNNNNNNNNN | 1 |
| C | 8547 hCoV-19/Spain/NC-IBV-001369/2020\|EPI_ISL_452532\|20 | ···················· | ···················· | ···················· | ···················· | NNNNNNNNNNNNNNNNNNNN | NNNNNNNNNNNNNNNNNNNN | 1 |
| C | 8550 hCoV-19/Spain/NC-IBV-001363/2020\|EPI_ISL_452538\|20 | ···················· | ···················· | ···················· | ···················· | NNNNNNNNNNNNNNNNNNNN | NNNNNNNNNNNNNNNNNNNN | 1 |
| C | 8553 hCoV-19/Spain/NC-IBV-001360/2020\|EPI_ISL_452541\|20 | ···················· | ···················· | ···················· | ···················· | ··NNNNN············· | ···················· | 1 |
| C | 8554 hCoV-19/Spain/AN-IBV-001329/2020\|EPI_ISL_452544\|20 | ···················· | ···················· | ···················· | ···················· | NNNNNNNNNNNNNNNNNNNN | ···················· | 1 |
| C | 8555 hCoV-19/Spain/AN-IBV-001327/2020\|EPI_ISL_452546\|20 | ···················· | ···················· | ···················· | ···················· | NNNNNNNNNNNNNNNNNNNN | ···················· | 1 |
| C | 8556 hCoV-19/Spain/AN-IBV-001326/2020\|EPI_ISL_452547\|20 | ···················· | ···················· | ···················· | ···················· | NNNNNNNNNNNNNNNNNNNN | NNNNNNNNNNNNNNNNNNNN | 1 |
| C | 8557 hCoV-19/Spain/AN-IBV-001324/2020\|EPI_ISL_452549\|20 | ···················· | ···················· | ···················· | ···················· | NNNNNNNNNNNNNNNNNNNN | NNNNNNNNNNNNNNNNN··· | 1 |
| C | 8558 hCoV-19/Spain/AN-IBV-001322/2020\|EPI_ISL_452551\|20 | ···················· | ···················· | ···················· | ···················· | NNNNNNNNNNNNNNNNNNNN | NNNNNNNNNNNNNNN····· | 1 |
| C | 8559 hCoV-19/Spain/AN-IBV-001321/2020\|EPI_ISL_452552\|20 | ···················· | ···················· | ···················· | ···················· | NNNNNNNNNNNNNNNNNNNN | NNNNNNNNNNNNNNNNNNNN | 1 |
| C | 8560 hCoV-19/Spain/AN-IBV-001194/2020\|EPI_ISL_452564\|20 | ···················· | ···················· | ···················· | ···················· | NNNNNNNNNNNNNNNNNNNN | NNNNNNNNNNNNNNNNNNNN | 1 |
| C | 8561 hCoV-19/Spain/AN-IBV-001191/2020\|EPI_ISL_452566\|20 | ···················· | ···················· | ···················· | ···················· | NNNNNNNNNNNNNNNNNNNN | NNNNNNNNNNNNNNN····· | 1 |
| C | 8563 hCoV-19/Spain/AN-IBV-001161/2020\|EPI_ISL_452575\|20 | ···················· | ···················· | ···················· | ···················· | NNNNNNNNNNNNNNNNNNNN | NNNNNNNNNNNNNNNNNNNN | 1 |
| C | 8564 hCoV-19/Spain/AN-IBV-001159/2020\|EPI_ISL_452576\|20 | ···················· | ···················· | ···················· | ···················· | NNNNNNNNNNNNNNNNNNNN | NNNNNNNNNNNNNNNNNNNN | 1 |
| C | 8566 hCoV-19/Spain/AN-IBV-001147/2020\|EPI_ISL_452578\|20 | ···················· | ···················· | ···················· | ···················· | NNNNNNNNNNNNNNNNNNNN | NNNNNNNNNNNNNNNNNNNN | 1 |
| C | 8567 hCoV-19/Spain/AN-IBV-001122/2020\|EPI_ISL_452579\|20 | ···················· | ···················· | ···················· | ···················· | NNNNNNNNNNNNNNNNNNNN | NNNNNNNNNNNNNNNNNNNN | 1 |
| C | 8568 hCoV-19/Spain/AN-IBV-001121/2020\|EPI_ISL_452580\|20 | ···················· | ···················· | ···················· | ···················· | NNNNNNNNNNNNNNNNNNNN | NNNNNNNNNNNNNNNNNNNN | 1 |
| C | 8569 hCoV-19/Spain/AN-IBV-001120/2020\|EPI_ISL_452581\|20 | ···················· | ···················· | ···················· | ···················· | NNNNNNNNNNNNNNNNNNNN | NNNNNNNNNNNNNNNNNNNN | 1 |
| C | 8570 hCoV-19/Spain/AN-IBV-001119/2020\|EPI_ISL_452582\|20 | ···················· | ···················· | ···················· | ···················· | NNNNNNNNNNNNNNNNNNNN | NNNNNNNNNNNNNNNNNNNN | 1 |
| C | 8571 hCoV-19/Spain/AN-IBV-001118/2020\|EPI_ISL_452583\|20 | ···················· | ···················· | ···················· | ···················· | NNNNNNNNNNNNNNNNNNNN | NNNNNNNNNNNNNNNNNNNN | 1 |
| C | 8572 hCoV-19/Spain/AN-IBV-001117/2020\|EPI_ISL_452584\|20 | ···················· | ···················· | ···················· | ···················· | NNNNNNNNNNNNNNNNNNNN | NNNNNNNNNNNNNNNNNNNN | 1 |
| C | 8574 hCoV-19/Spain/AN-IBV-001099/2020\|EPI_ISL_452588\|20 | ···················· | ···················· | ···················· | ···················· | NNNNNNNNNNNNNNNNNNNN | ···················· | 1 |
| C | 8575 hCoV-19/Spain/AN-IBV-001097/2020\|EPI_ISL_452590\|20 | ···················· | ···················· | ···················· | ···················· | ·NNNNNNNNNNNNNNNNNNN | NNNNNNNNNNNNNNNNNNNN | 1 |
| C | 8576 hCoV-19/Spain/AN-IBV-001094/2020\|EPI_ISL_452592\|20 | ···················· | ···················· | ···················· | ···················· | NNNNNNNNNNNNNNNNNNNN | NNNNNNNNNNNNNNNNNNNN | 1 |
| C | 8578 hCoV-19/Spain/AN-IBV-001092/2020\|EPI_ISL_452594\|20 | ···················· | ···················· | ···················· | ···················· | NNNNNNNNNNNNNNNNNNNN | NNNNNNNNNNNNNNNNNNNN | 1 |
| C | 8579 hCoV-19/Spain/AN-IBV-001091/2020\|EPI_ISL_452595\|20 | ···················· | ···················· | ···················· | ···················· | NNNNNNNNNNNNNNNNNNNN | NNNNNNNNNNNNNNNNNNNN | 1 |
| C | 8580 hCoV-19/Spain/AN-IBV-001090/2020\|EPI_ISL_452596\|20 | ···················· | ···················· | ···················· | ···················· | NNNNNNNNNNNNNNNNNNNN | NNNNNNNNNNNNNNNNNNNN | 1 |
| C | 8734 hCoV-19/Spain/NC-IBV-001401/2020\|EPI_ISL_452500\|20 | ···················· | ···················· | ···················· | ···················· | NNNNNNNNNNNNNNNNNNNN | NNNNNNNNNNNNNNNNNNNN | 1 |
| C | 8736 hCoV-19/Spain/NC-IBV-001396/2020\|EPI_ISL_452505\|20 | ···················· | ···················· | ···················· | ···················· | NNNNNNNNNNNNNNNNNNNN | NNNNNNNNNNNNNNNNNNNN | 1 |
| C | 8737 hCoV-19/Spain/NC-IBV-001395/2020\|EPI_ISL_452506\|20 | ···················· | ···················· | ···················· | ···················· | NNNNNNNNNNNNNNNNNNNN | NNNNNNNNNNNNNNNNNNNN | 1 |
| C | 8738 hCoV-19/Spain/NC-IBV-001394/2020\|EPI_ISL_452507\|20 | ···················· | ···················· | ···················· | ···················· | NNNNNNNNNNNNNNNNNNNN | ···················· | 1 |
| C | 8740 hCoV-19/Spain/RI-IBV-001392/2020\|EPI_ISL_452509\|20 | ···················· | ···················· | ···················· | ···················· | NNNNNNNNNNNNNNNNNNNN | NNNNNNNNNNNNNNNNNNNN | 1 |
| C | 8741 hCoV-19/Spain/AN-IBV-001320/2020\|EPI_ISL_452553\|20 | ···················· | ···················· | ···················· | ···················· | NNNNNNN············· | ···················· | 1 |
| C | 8742 hCoV-19/Spain/AN-IBV-001272/2020\|EPI_ISL_452554\|20 | ···················· | ···················· | ···················· | ···················· | NNNNNNNNNNNNNNNNNNNN | NNNNNNNNNNNNNNNNNNNN | 1 |
| C | 8743 hCoV-19/Spain/AN-IBV-001271/2020\|EPI_ISL_452555\|20 | ···················· | ···················· | ···················· | ···················· | NNNNNNNNNNNNNNNNNNNN | NNNNNNNNNNNNNNN····· | 1 |
| C | 8746 hCoV-19/Spain/AN-IBV-001214/2020\|EPI_ISL_452558\|20 | ···················· | ···················· | ···················· | ···················· | NNNNNNNNNNNNNNNNNNNN | NNNNNNNNNNNNNNNNNNNN | 1 |
| C | 8747 hCoV-19/Spain/AN-IBV-001213/2020\|EPI_ISL_452559\|20 | ···················· | ···················· | ···················· | ···················· | NNNNNNNNNNNNNNNNNNNN | NNNNNNNNNNNNNNNNNNNN | 1 |
| C | 8748 hCoV-19/Spain/AN-IBV-001211/2020\|EPI_ISL_452560\|20 | ···················· | ···················· | ···················· | ···················· | NNNNNNNNNNNNNNNNNNNN | NNNNNNNNNNNNNNNNNNNN | 1 |
| C | 8750 hCoV-19/Spain/AN-IBV-001196/2020\|EPI_ISL_452562\|20 | ···················· | ···················· | ···················· | ···················· | ·NNNNNN············· | ···················· | 1 |
| C | 8752 hCoV-19/Spain/AN-IBV-001168/2020\|EPI_ISL_452571\|20 | ···················· | ···················· | ···················· | ···················· | NNNNNNNNNNNNNNNNNNNN | NNNNNNNNNNNNNNNNNNNN | 1 |
| C | 8753 hCoV-19/Spain/AN-IBV-001164/2020\|EPI_ISL_452572\|20 | ···················· | ···················· | ···················· | ···················· | ·NNNNNN············· | ···················· | 1 |
| C | 8754 hCoV-19/Spain/AN-IBV-001163/2020\|EPI_ISL_452573\|20 | ···················· | ···················· | ···················· | ···················· | NNNNNNNNNNNNNNNNNNNN | ···················· | 1 |
| C | 8755 hCoV-19/Spain/AN-IBV-001162/2020\|EPI_ISL_452574\|20 | ···················· | ···················· | ···················· | ···················· | NNNNNNNNNNNNNNNNNNNN | NNNNNNNNNNNNNNN····· | 1 |
| C | 9066 hCoV-19/England/NORW-EA27F/2020\|EPI_ISL_457336\|202 | ···················· | ···················· | ···················· | ···················· | NNNNNNNNNNNNNNNNNNNN | NNNNNNNNNNNNNNNNNNNN | 1 |
| C | 9067 hCoV-19/England/NORW-EA28E/2020\|EPI_ISL_457337\|202 | ···················· | ···················A | ···················· | ···················· | NNNN················ | ···················· | 1 |
| C | 9068 hCoV-19/England/NORW-EA29D/2020\|EPI_ISL_457338\|202 | ···················· | ···················· | ···················· | ···················· | NNNNNNNNNNNNNN······ | ···················· | 1 |
| C | 9077 hCoV-19/England/NORW-EAA77/2020\|EPI_ISL_457350\|202 | ···················· | ···················· | ···················· | ···················· | ···················· | ·NNNN··············· | 1 |
| C | 9078 hCoV-19/England/NORW-EAA95/2020\|EPI_ISL_457351\|202 | ···················· | ···················· | ···················· | ···················· | NNNNNNNNNNNNNNNNNNNN | NNNNNNNNNNNNNNNNNNNN | 1 |
| C | 9080 hCoV-19/England/NORW-EAAB3/2020\|EPI_ISL_457353\|202 | ···················· | ···················· | ···················· | ···················· | NNNN················ | ···················· | 1 |
| C | 9090 hCoV-19/England/NORW-EAC71/2020\|EPI_ISL_457366\|202 | ···················· | ···················· | ···················· | ···················· | NNNNNNNNNNNNNNNNNNNN | ···················· | 1 |
| C | 9220 hCoV-19/England/NORW-E712A/2020\|EPI_ISL_449004\|202 | ···················· | ···················· | ···················· | ···················· | NNNNNNNNNNNNNNNNNNNN | NNNNNNNNNNNNNNN····· | 1 |
| C | 9231 hCoV-19/England/NORW-E72AF/2020\|EPI_ISL_449017\|202 | ···················· | ···················· | ···················· | ···················· | NNNNNNNNNNNNNNNNNNNN | NNNNNNNNNNNNNNNNNNNN | 1 |
| C | 9240 hCoV-19/England/NORW-E7360/2020\|EPI_ISL_449027\|202 | ···················· | ···················· | ···················· | ···················· | NNNNNNNNNNNN········ | NNNN················ | 1 |
| C | 9259 hCoV-19/England/NORW-E7D62/2020\|EPI_ISL_449089\|202 | ···················· | ···················· | ···················· | ···················· | NNN················· | NNNN·NNNN··········· | 1 |
| C | 9342 hCoV-19/Spain/PV-IBV-000942/2020\|EPI_ISL_452630\|20 | ···················· | ···················· | ···················· | ···················· | NNNNNNNNNNNNNNNNNNNN | NNNNNNNNNNNNNNNNNNNN | 1 |
| C | 9344 hCoV-19/Spain/PV-IBV-000940/2020\|EPI_ISL_452632\|20 | ···················· | ···················· | ···················· | ···················· | NNNNNNNNNNNNNNNNNNNN | NNNNNNNNNNNNNNNNNNNN | 1 |

| | | | | | | | | |
|---|---|---|---|---|---|---|---|---|
| C | 9345 hCoV-19/Spain/PV-IBV-000939/2020\|EPI_ISL_452633\|20 | ···················· | ···················· | ···················· | ···················· | NNNNNNNNNNNN········ | ···················· | 1 |
| C | 9346 hCoV-19/Spain/PV-IBV-000938/2020\|EPI_ISL_452634\|20 | ···················· | ···················· | ···················· | ···················· | NNNNNNNNNNNNNNNNNNNN | NNNNNNNNNNNNNNNNNNNN | 1 |
| C | 9347 hCoV-19/Spain/PV-IBV-000937/2020\|EPI_ISL_452635\|20 | ···················· | ···················· | ···················· | ···················· | NNNNNNNNNNNNNNNNNNNN | NNNNNNNNNNNNNNNNNNNN | 1 |
| C | 9348 hCoV-19/Spain/PV-IBV-000936/2020\|EPI_ISL_452636\|20 | ···················· | ···················· | ···················· | ···················· | NNNNNNNNNNNNNNNNNNNN | ··············N····· | 1 |
| C | 9350 hCoV-19/Spain/PV-IBV-000934/2020\|EPI_ISL_452638\|20 | ···················· | ···················· | ···················· | ···················· | NNNNNNNNN··········· | ···················· | 1 |
| C | 9351 hCoV-19/Spain/PV-IBV-000933/2020\|EPI_ISL_452639\|20 | ···················· | ···················· | ···················· | ···················· | NNNNNNNNNNNNNNNNNNNN | NNNNNNNNNNNNNNNN···N | 1 |
| C | 9352 hCoV-19/Spain/PV-IBV-000932/2020\|EPI_ISL_452640\|20 | ···················· | ···················· | ···················· | ···················· | NNNNNNN············· | ···················· | 1 |
| C | 9353 hCoV-19/Spain/PV-IBV-000931/2020\|EPI_ISL_452641\|20 | ···················· | ···················· | ···················· | ···················· | NNNNNNNNNNNNNNNNNNNN | ···················· | 1 |
| C | 9354 hCoV-19/Spain/PV-IBV-000929/2020\|EPI_ISL_452642\|20 | ···················· | ···················· | ···················· | ···················· | NNNNNNNNNNNNNNNNNNNN | NNNNNNNNNNNNNNNNNNNN | 1 |
| C | 9355 hCoV-19/Spain/PV-IBV-000928/2020\|EPI_ISL_452643\|20 | ···················· | ···················· | ···················· | ···················· | NNNNNNNNNNNNNNNNNNNN | ···················· | 1 |
| C | 9356 hCoV-19/Spain/PV-IBV-000927/2020\|EPI_ISL_452644\|20 | ···················· | ···················· | ···················· | ···················· | NNNNNNNNNNNN········ | ···················· | 1 |
| C | 9357 hCoV-19/Spain/PV-IBV-000926/2020\|EPI_ISL_452645\|20 | ···················· | ···················· | ···················· | ···················· | NNNNNNNNNNNNNNNNNNNN | NNNNNNNNNNNNNNNNNNNN | 1 |
| C | 9358 hCoV-19/Spain/PV-IBV-000925/2020\|EPI_ISL_452646\|20 | ···················· | ···················· | ···················· | ···················· | NNNNNNNNNNNNNNNNNNNN | NNNNNNNNNNNNNNNNNNNN | 1 |
| C | 9360 hCoV-19/Spain/PV-IBV-000923/2020\|EPI_ISL_452648\|20 | ···················· | ···················· | ···················· | ···················· | NNNNNNNNNNNNNNNNNNNN | ···········NNNN····· | 1 |
| C | 9361 hCoV-19/Spain/PV-IBV-000922/2020\|EPI_ISL_452649\|20 | ···················· | ···················· | ···················· | ···················· | NNNNNNNNNNNNNNNNNNNN | NNNNNNNNNNNNNNNNNNNN | 1 |
| C | 9362 hCoV-19/Spain/PV-IBV-000920/2020\|EPI_ISL_452650\|20 | ···················· | ···················· | ···················· | ···················· | NNNNNNNNNNNNNNNNNNNN | ···················· | 1 |
| C | 9364 hCoV-19/Spain/PV-IBV-000917/2020\|EPI_ISL_452652\|20 | ···················· | ···················· | ···················· | ···················· | NNNNNNNNNNNNNNNNNNNN | NNNNNNNNNNNNNNNNNNNN | 1 |
| C | 9365 hCoV-19/Spain/PV-IBV-000915/2020\|EPI_ISL_452653\|20 | ···················· | ···················· | ···················· | ···················· | NNNNNNNNNNNNNNNNNNNN | NNNNNNNNNNNNNNNNNNNN | 1 |
| C | 9366 hCoV-19/Spain/PV-IBV-000910/2020\|EPI_ISL_452657\|20 | ···················· | ···················· | ···················· | ···················· | ··NNNNN············· | ···················· | 1 |
| C | 9367 hCoV-19/Spain/PV-IBV-000909/2020\|EPI_ISL_452658\|20 | ···················· | ···················· | ···················· | ···················· | NNNNNNN············· | ···················· | 1 |
| C | 9368 hCoV-19/Spain/PV-IBV-000908/2020\|EPI_ISL_452659\|20 | ···················· | ···················· | ···················· | ···················· | NNNNNNNNNNNNNNNNNNNN | NNNNNNNNNNNNNNNNNNNN | 1 |
| C | 9369 hCoV-19/Spain/PV-IBV-000906/2020\|EPI_ISL_452661\|20 | ···················· | ···················· | ···················· | ···················· | NNNNNNNNNNNNNNNNNNNN | NNNNNNNNNNNNNNNNNNNN | 1 |
| C | 9370 hCoV-19/Spain/PV-IBV-000905/2020\|EPI_ISL_452662\|20 | ···················· | ···················· | ···················· | ···················· | NNNNNNNNNNNNNNNNNNNN | NNNNNNNNNNNNNNNNNNNN | 1 |
| C | 9371 hCoV-19/Spain/PV-IBV-000904/2020\|EPI_ISL_452663\|20 | ···················· | ···················· | ···················· | ···················· | NNNNNNNNNNNNNNNNNNNN | ···················· | 1 |
| C | 9372 hCoV-19/Spain/PV-IBV-000902/2020\|EPI_ISL_452664\|20 | ···················· | ···················· | ···················· | ···················· | NNNNNNNNNNNNNNNNNNNN | NNNNNNNNNNNNNNNNNNNN | 1 |
| C | 9373 hCoV-19/Spain/PV-IBV-000901/2020\|EPI_ISL_452665\|20 | ···················· | ···················· | ···················· | ···················· | NNNNNNNNNNNNNNNNNNNN | NNNNNNNNNNNNNNNN···· | 1 |
| C | 9374 hCoV-19/Spain/PV-IBV-000897/2020\|EPI_ISL_452666\|20 | ···················· | ···················· | ···················· | ···················· | NNNNNNNNNNNNNNNNNNNN | NNNNNNNNNNNNNNNNNNNN | 1 |
| C | 9375 hCoV-19/Spain/PV-IBV-000894/2020\|EPI_ISL_452669\|20 | ···················· | ···················· | ···················· | ···················· | NNNNNNNNNNNNNNNNNNNN | NNNNNNNNNNNNNNNN···· | 1 |
| C | 9376 hCoV-19/Spain/PV-IBV-000893/2020\|EPI_ISL_452670\|20 | ···················· | ···················· | ···················· | ···················· | NNNNNNNNNNNNNNNNNNNN | ···················· | 1 |
| C | 9378 hCoV-19/Spain/PV-IBV-000891/2020\|EPI_ISL_452672\|20 | ···················· | ···················· | ···················· | ···················· | NNNNNNNNNNNNNNNNNNNN | ···················· | 1 |
| C | 9498 hCoV-19/England/NORW-E7755/2020\|EPI_ISL_449068\|202 | ···················· | ···················· | ···················· | ···················· | NNNNNNNNNNNNNNNNNNNN | NNNNNNNNNNNNNNNNNNNN | 1 |
| C | 9549 hCoV-19/Spain/AN-IBV-001086/2020\|EPI_ISL_452600\|20 | ···················· | ···················· | ···················· | ···················· | NNNNNNN···NNNNNNNNNN | NNNNNNNNNNNNNNNN···· | 1 |
| C | 9551 hCoV-19/Spain/AN-IBV-001083/2020\|EPI_ISL_452603\|20 | ···················· | ···················· | ···················· | ···················· | NNNNNNNNNNNNNNNNNNNN | NNNNNNNNNNNNNNNNNNNN | 1 |
| C | 9552 hCoV-19/Spain/AN-IBV-001081/2020\|EPI_ISL_452604\|20 | ···················· | ···················· | ···················· | ···················· | NNNNNNNNNNNNNNNNNNNN | NNNNNNNNNNNNNNNNNNNN | 1 |
| C | 9553 hCoV-19/Spain/AN-IBV-001080/2020\|EPI_ISL_452605\|20 | ···················· | ···················· | ···················· | ···················· | NNNNNNN············· | ···················· | 1 |
| C | 9555 hCoV-19/Spain/AN-IBV-001075/2020\|EPI_ISL_452609\|20 | ···················· | ···················· | ···················· | ···················· | NNNNNNNNNNNNNNNNNNNN | ···················· | 1 |
| C | 9556 hCoV-19/Spain/AN-IBV-001066/2020\|EPI_ISL_452610\|20 | ···················· | ···················· | ···················· | ···················· | NNNNNNNNNNNNNNNNNNNN | NNNNNNNNNNNNNNNNNNNN | 1 |
| C | 9557 hCoV-19/Spain/AN-IBV-001064/2020\|EPI_ISL_452612\|20 | ···················· | ···················· | ···················· | ···················· | NNNNNNNNNNNNNNNNNNNN | NNNNNNNNNNNNNNNNNNNN | 1 |
| C | 9558 hCoV-19/Spain/AN-IBV-001060/2020\|EPI_ISL_452613\|20 | ···················· | ···················· | ···················· | ···················· | NNNNNNNNNNNNNNNNNNNN | NNNNNNNNNNNNNNNNNNNN | 1 |
| C | 9559 hCoV-19/Spain/AN-IBV-001057/2020\|EPI_ISL_452615\|20 | ···················· | ···················· | ···················· | ···················· | NNNNNNNNNNNNNNNNNNNN | NNNNNNNNNNNNNNNNNNNN | 1 |
| C | 9560 hCoV-19/Spain/PV-IBV-000957/2020\|EPI_ISL_452618\|20 | ···················· | ···················· | ···················· | ···················· | NNNNNNNNNNNNNNNNNNNN | NNNNNNNNNNNNNNNNNNNN | 1 |
| C | 9562 hCoV-19/Spain/PV-IBV-000954/2020\|EPI_ISL_452621\|20 | ···················· | ···················· | ···················· | ···················· | NNNNNNNNNNNNNNNNNNNN | ·········NNNNNN····· | 1 |
| C | 9563 hCoV-19/Spain/PV-IBV-000953/2020\|EPI_ISL_452622\|20 | ···················· | NNNNNNNNNNNNNNNNNNNN | ···················· | ···················· | NNNNNNN············· | ···················· | 1 |
| C | 9564 hCoV-19/Spain/PV-IBV-000950/2020\|EPI_ISL_452623\|20 | ···················· | ···················· | ···················· | ···················· | NNNNNNNNNNNNNNNNNNNN | NNNNNNNNNNNNNNNNNNNN | 1 |
| C | 9565 hCoV-19/Spain/PV-IBV-000949/2020\|EPI_ISL_452624\|20 | ···················· | ···················· | ···················· | ···················· | NNNNNNNNNNNNNNNNNNNN | NNNNNNNNNNNNNNNNNNNN | 1 |
| C | 9566 hCoV-19/Spain/PV-IBV-000948/2020\|EPI_ISL_452625\|20 | ···················· | ···················· | ···················· | ···················· | NNNNNNNNNNNNNNNNNNNN | NNNNNNNNNNNNNNNNNNNN | 1 |
| C | 9567 hCoV-19/Spain/PV-IBV-000947/2020\|EPI_ISL_452626\|20 | ···················· | ···················· | ···················· | ···················· | NNNNNNNNNNNNNNNNNNNN | ···················· | 1 |
| C | 9568 hCoV-19/Spain/PV-IBV-000946/2020\|EPI_ISL_452627\|20 | ···················· | ···················· | ···················· | ···················· | NNNNNNNNNNNNNNNNNNNN | ···················· | 1 |
| C | 9569 hCoV-19/Spain/PV-IBV-000887/2020\|EPI_ISL_452675\|20 | ···················· | ···················· | ···················· | ···················· | NNNNNNNNNNNNNNNNNNNN | ···················· | 1 |
| C | 9571 hCoV-19/Spain/PV-IBV-000881/2020\|EPI_ISL_452678\|20 | ···················· | ···················· | ···················· | ···················· | NNNNNNNNNNNNNNNNNNNN | ···················· | 1 |
| C | 9572 hCoV-19/Spain/PV-IBV-000880/2020\|EPI_ISL_452679\|20 | ···················· | ···················· | ···················· | ···················· | NNNNNNNNNNNNNNNNNNNN | ···················· | 1 |
| C | 9573 hCoV-19/Spain/PV-IBV-000879/2020\|EPI_ISL_452680\|20 | ···················· | ···················· | ···················· | ···················· | NNNNNNN·N··········· | ···················· | 1 |
| C | 9574 hCoV-19/Spain/PV-IBV-000876/2020\|EPI_ISL_452681\|20 | ···················· | ···················· | ···················· | ···················· | NNNNNNNNNNNNNNNNNNNN | NNNNNNNNNNNNNNNNNNNN | 1 |
| C | 9578 hCoV-19/Spain/PV-IBV-000869/2020\|EPI_ISL_452686\|20 | ···················· | ···················· | ···················· | ···················· | NNNNNNNNNNNN········ | NNNNNNNNNNNNNNNN···· | 1 |
| C | 9579 hCoV-19/Spain/PV-IBV-000867/2020\|EPI_ISL_452688\|20 | ···················· | ···················· | ···················· | ···················· | ·NNNNNN············· | ···················· | 1 |
| C | 9580 hCoV-19/Spain/PV-IBV-000866/2020\|EPI_ISL_452689\|20 | ···················· | ···················· | ···················· | ···················· | NNNNNNNNNNNNNNNNNNNN | NNNNNNNNNNNNNNNN···· | 1 |
| C | 9581 hCoV-19/Spain/PV-IBV-000864/2020\|EPI_ISL_452691\|20 | ···················· | ···················· | ···················· | ···················· | NNNNNNNNNNNNNNNNNNNN | NNNNNNNNNNNNNNNNNNNN | 1 |
| C | 9582 hCoV-19/Spain/PV-IBV-000860/2020\|EPI_ISL_452692\|20 | ···················· | ···················· | ···················· | ···················· | NNNNNNNNNNNN····NNN· | NNNNNNNNNNNNNNNN···· | 1 |
| C | 9583 hCoV-19/Spain/PV-IBV-000859/2020\|EPI_ISL_452693\|20 | ···················· | ···················· | ···················· | ···················· | NNNNNNNNNNNNNNNNNNNN | NNNNNNNNNNNNNNNNNNNN | 1 |
| C | 9584 hCoV-19/Spain/PV-IBV-000858/2020\|EPI_ISL_452694\|20 | ···················· | ···················· | ···················· | ···················· | NNNNNNNNNNNNNNNNNNNN | NNNNNNNNNNNNNNNNNNNN | 1 |
| C | 9585 hCoV-19/Spain/PV-IBV-000857/2020\|EPI_ISL_452695\|20 | ···················· | ···················· | ···················· | ···················· | ·NNNNNN············· | ···················· | 1 |
| C | 9586 hCoV-19/Spain/PV-IBV-000856/2020\|EPI_ISL_452696\|20 | ···················· | ···················· | ···················· | ···················· | NNNNNNNNNNNNNNNNNNNN | NNNNNNNNNNNNNNNNNNNN | 1 |
| C | 9587 hCoV-19/Spain/PV-IBV-000853/2020\|EPI_ISL_452698\|20 | ···················· | ···················· | ···················· | ···················· | NNNNNNN············· | ···················· | 1 |
| C | 9588 hCoV-19/Spain/PV-IBV-000852/2020\|EPI_ISL_452699\|20 | ···················· | ···················· | ···················· | ···················· | NNNNNNNNNNNNNNNNNNNN | NNNNNNNNNNNNNNNN···· | 1 |
| C | 9666 hCoV-19/England/NORW-EB42D/2020\|EPI_ISL_457430\|202 | ···················· | ···················· | ···················· | ···················· | NNNNNNNNNNNNNNNNNNNN | NNNNNNNNNNNNNNNNNNNN | 1 |
| C | 9729 hCoV-19/Colombia/ANT-INS-79253/2020\|EPI_ISL_456117 | ···················· | ···················· | ···················· | ···················· | NNNN················ | ···················· | 1 |
| C | 9730 hCoV-19/Colombia/ANT-INS-80825/2020\|EPI_ISL_456119 | ···················· | ···················· | ···················· | ···················· | ·NNNNNNNNNNNNNNNNNNN | ···················· | 1 |
| C | 9731 hCoV-19/Colombia/DC-INS-79190/2020\|EPI_ISL_456120\| | ···················· | ···················· | ···················· | ···················· | NNNNNNNNNNNNNNNNNNNN | NNNNNNNNNNNNNNNNNNNN | 1 |
| C | 9732 hCoV-19/Colombia/NSA-INS-80656/2020\|EPI_ISL_456126 | ···················· | ···················· | ···················· | ···················· | NNNNNNNNNNNNNNNNNNNN | NNNNNNNNNNNNNNNNNNNN | 1 |
| C | 9765 hCoV-19/England/NORW-EB663/2020\|EPI_ISL_457452\|202 | ···················· | ···················· | ···················· | ···················· | NNNNNNNNNNNNNNNNN··· | N··················· | 1 |
| D | 7 hCoV-19/England/20149001204/2020\|EPI_ISL_466082\|20 | ···················· | ···················· | ···················· | ···················· | ··NNNNNNNNNNNNNNNNNN | ···················· | 1 |
| D | 15 hCoV-19/England/20149003704/2020\|EPI_ISL_466095\|20 | ···················· | ···················· | ···················· | ···················· | ··N················· | NNNNNNNNNNNNNNNNNNNN | 1 |
| D | 16 hCoV-19/England/20149003804/2020\|EPI_ISL_466096\|20 | ···················· | ···················· | ···················· | ···················· | NNNNNNNNNNNNNNNNNNNN | NNNNNNNNNNNNNNNNNNNN | 1 |
| D | 17 hCoV-19/England/20149003904/2020\|EPI_ISL_466097\|20 | ···················· | ···················· | ···················· | ···················· | NNNNNNNNNNNNNNNNNNNN | NNNNNNNNNNNNNNNNNNNN | 1 |
| D | 20 hCoV-19/England/20149004404/2020\|EPI_ISL_466101\|20 | ···················· | ···················· | ···················· | ···················· | ···················· | NNNN················ | 1 |
| D | 21 hCoV-19/England/20149004504/2020\|EPI_ISL_466102\|20 | ···················· | ···················· | ···················· | ···················· | NNNNNNNNNNNNNNNNNNNN | ···········N········ | 1 |
| D | 87 hCoV-19/Belgium/UGent-79/2020\|EPI_ISL_468729\|2020- | ···················· | ···················· | ···················· | ···················· | NNNNNNNNNNNNNNNNNNNN | NNNNNNNNNNNNNNNNNNNN | 1 |
| D | 88 hCoV-19/Belgium/UGent-78/2020\|EPI_ISL_468730\|2020- | ···················· | ···················· | ···················· | ···················· | NNNNNNNNNNNNNNNNNNNN | NNNNNNNNNNNNNNNNNNNN | 1 |
| D | 117 hCoV-19/Spain/AR-IBV-2466/2020\|EPI_ISL_468765\|2020 | ···················· | ···················· | ···················· | ···················· | ·NNNNNNNNNNNNNNNNNNN | NNNNNNNNNNNNNNNN···· | 1 |
| D | 118 hCoV-19/Spain/AR-IBV-2468/2020\|EPI_ISL_468766\|2020 | ···················· | ···················· | ···················· | ···················· | NNNNNNNNNNNNNNNNNNNN | NNNNNNNNNNNNNNNNNNNN | 1 |

| | | | | | | | | |
|---|---|---|---|---|---|---|---|---|
| D | 119 hCoV-19/Spain/AR-IBV-2416/2020\|EPI_ISL_468768\|2020 | ···················· | ···················· | ···················· | ···················· | NNNNNNN············· | ···················· | 1 |
| D | 120 hCoV-19/Spain/AR-IBV-2476/2020\|EPI_ISL_468769\|2020 | ···················· | ···················· | ···················· | ···················· | NNNNNNN············· | ···················· | 1 |
| D | 121 hCoV-19/Spain/AR-IBV-2451/2020\|EPI_ISL_468770\|2020 | ···················· | ···················· | ···················· | ···················· | NNNNNNNNNNNNNNNNNNNN | NNNNNNNNNNNNNNNNNNNN | 1 |
| D | 123 hCoV-19/Spain/AR-IBV-2414/2020\|EPI_ISL_468772\|2020 | ···················· | ···················· | ···················· | ···················· | NNNNNNN············· | ···················· | 1 |
| D | 124 hCoV-19/Spain/AR-IBV-2470/2020\|EPI_ISL_468773\|2020 | ···················· | ···················· | ···················· | ···················· | NNNNNNNNNNNNNNNNNNNN | ···················· | 1 |
| D | 125 hCoV-19/Spain/AR-IBV-2454/2020\|EPI_ISL_468774\|2020 | ···················· | ···················· | ···················· | ···················· | NNNNNNNNNNNNNNNNNNNN | NNNNNNNNNNNNNNNN···· | 1 |
| D | 126 hCoV-19/Spain/AR-IBV-2475/2020\|EPI_ISL_468775\|2020 | ···················· | ···················· | ···················· | ···················· | NNNNNNNNNNNNNNNNNNNN | NNNNNNNNNNNNNNNNNNNN | 1 |
| D | 127 hCoV-19/Spain/AR-IBV-2442/2020\|EPI_ISL_468776\|2020 | ···················· | ···················· | ···················· | ···················· | NNNNNNNNNNNNNNNNNNNN | NNNNNNNNNNNNNNNNNNNN | 1 |
| D | 250 hCoV-19/Scotland/CVR3458/2020\|EPI_ISL_478064\|2020- | ···················· | ···················A | ···················· | ···················· | NNNNNNNNNNNNNNNNNNNN | ···················· | 1 |
| D | 261 hCoV-19/England/20144063304/2020\|EPI_ISL_466004\|20 | ···················· | ···················· | ···················· | ···················· | ·NNNNNNNNNNNNNNNNNNN | NNNNNNNNNNNNNNNNNNNN | 1 |
| D | 262 hCoV-19/England/20144067304/2020\|EPI_ISL_466005\|20 | ···················· | ···················· | ···················· | ···················· | ·NNNNNNNNNNNNNNNNNNN | NNNNNNNNNNNNNNNNNNNN | 1 |
| D | 264 hCoV-19/England/20144075304/2020\|EPI_ISL_466008\|20 | ···················· | ···················· | ···················· | ···················· | ·NN··NNN···NNNNNNNNN | NNNNNNNNNNNNNNNNNNNN | 1 |
| D | 274 hCoV-19/England/20146013704/2020\|EPI_ISL_466029\|20 | ···················· | ···················· | ···················· | ···················· | ·····NNNNNNNNNNNNNNN | NNNNNNNNNNNNNNNNNNNN | 1 |
| D | 287 hCoV-19/England/20146061104/2020\|EPI_ISL_466045\|20 | ···················· | ···················· | ···················· | ···················· | ··NNNNNNNNNNNNNNNNNN | NNNNNNNNNNNNNNNNNNNN | 1 |
| D | 291 hCoV-19/England/20148002204/2020\|EPI_ISL_466050\|20 | ···················· | ···················· | ···················· | ···················· | ·NNNNNNNNNNNNNNNNNNN | NNNNNNNNNNNNNNNNNNNN | 1 |
| D | 292 hCoV-19/England/20148003504/2020\|EPI_ISL_466051\|20 | ···················· | ···················· | ···················· | ···················· | ·NNNNNNNNNNNNNNNNNNN | NNNNNNNNNNNNNNNNNNNN | 1 |
| D | 295 hCoV-19/England/20148033504/2020\|EPI_ISL_466057\|20 | ···················· | ···················· | ···················· | ···················· | ·NNNNNNNNNNNNNNNNNNN | NNNNNNNNNNNNNNNNNNNN | 1 |
| D | 296 hCoV-19/England/20148034104/2020\|EPI_ISL_466059\|20 | ···················· | ···················· | ···················· | ···················· | ·NNNNNNNNNNNNNNNNNNN | NNNNNNNNNNNNNNNN···· | 1 |
| D | 299 hCoV-19/England/20148035504/2020\|EPI_ISL_466063\|20 | ···················· | ···················· | ···················· | ···················· | ·NNNNNNNNNNNNNNNNNNN | NNNNNNNNNNNNNNNNNNNN | 1 |
| D | 301 hCoV-19/England/20148037904/2020\|EPI_ISL_466066\|20 | ···················· | ···················· | ···················· | ···················· | ···················· | NNNNNNNNN··NNNN····· | 1 |
| D | 306 hCoV-19/England/20148067004/2020\|EPI_ISL_466075\|20 | ···················· | ···················· | ···················· | ···················· | ···················· | NNNNNNNNNNNNNNNNNNNN | 1 |
| D | 372 hCoV-19/South Africa/KRISP-0170/2020\|EPI_ISL_46752 | ···················· | ···················· | ···················· | ···················· | ··NNNNNNNNNNNNNNNNNN | NNNNNNNNNNNNNNNNNNNN | 1 |
| D | 411 hCoV-19/Spain/AR-IBV-2424/2020\|EPI_ISL_468850\|2020 | ···················· | ···················· | ···················· | ···················· | NNNNNNNNNNNNNNNNNNNN | NNNNNNNNNNNNNNNN···· | 1 |
| D | 412 hCoV-19/Spain/AR-IBV-2396/2020\|EPI_ISL_468851\|2020 | ···················· | ···················· | ···················· | ···················· | ·NNNNNN······NNNNNNN | ···················· | 1 |
| D | 413 hCoV-19/Spain/AR-IBV-2452/2020\|EPI_ISL_468852\|2020 | ···················· | ···················· | ···················· | ···················· | NNNNNNNNNNNNNNNNNNNN | NNNNNNNNNNNNNNNNNNNN | 1 |
| D | 414 hCoV-19/Spain/AR-IBV-2395/2020\|EPI_ISL_468853\|2020 | ···················· | ···················· | ···················· | ···················· | NNNNNNNNNNNNNNNNNNNN | NNNNNNNNNNNNNNNNNNNN | 1 |
| D | 415 hCoV-19/Spain/AR-IBV-2462/2020\|EPI_ISL_468854\|2020 | ···················· | ···················· | ···················· | ···················· | ·NNNNNNNNNNNNNNNN··· | ···················· | 1 |
| D | 487 hCoV-19/Scotland/CVR3467/2020\|EPI_ISL_478070\|2020- | ···················· | ···················A | ···················· | ···················· | NNNNNNNNNNNNNNNNNNNN | ···················· | 1 |
| D | 509 hCoV-19/India/MH-NIV-15879/2020\|EPI_ISL_479503\|202 | ···················· | ···················· | ···················· | ·········NNNNNN····· | ···················· | ···················· | 1 |
| D | 512 hCoV-19/India/MH-NIV-16936/2020\|EPI_ISL_479506\|202 | ···················· | ···················· | ···················· | ·········NNNNNNNNNNN | ···················· | ···················· | 1 |
| D | 540 hCoV-19/England/20149049604/2020\|EPI_ISL_466204\|20 | ···················· | ···················· | ···················· | ···················· | ···················· | NNNNNNNNNNNNNNNNNNNN | 1 |
| D | 549 hCoV-19/England/20149054604/2020\|EPI_ISL_466218\|20 | ···················· | ···················· | ···················· | ···················· | ·NNNNNNNNNNNN··N·N·· | N·················NN | 1 |
| D | 552 hCoV-19/England/20150032804/2020\|EPI_ISL_466225\|20 | ··T················· | ···················· | ···················· | ···················· | NNNNNNNNNNNNNNNNNNNN | NNNNNNNNNNNNNNNNNNNN | 1 |
| D | 553 hCoV-19/England/20150033004/2020\|EPI_ISL_466226\|20 | ···················· | ···················· | ···················· | ···················· | ···················· | NNNNNNNNNNNNNNNNNNNN | 1 |
| D | 555 hCoV-19/England/20150040104/2020\|EPI_ISL_466230\|20 | ···················· | ···················· | ···················· | ···················· | ·NNNNNNNNNNNNNNNNNNN | NNNNNNNNNNNNNNNNNNNN | 1 |
| D | 587 hCoV-19/Spain/AR-IBV-2390/2020\|EPI_ISL_468800\|2020 | ···················· | ···················· | ···················· | ···················· | NNNNNNNNNNNNNNNNNNNN | NNNNNNNNNNNNNNNNNNNN | 1 |
| D | 588 hCoV-19/Spain/AR-IBV-2467/2020\|EPI_ISL_468801\|2020 | ···················· | ···················· | ···················· | ···················· | NNNNNNNNNNNNNNNNNNNN | NNNNNNNNNNNNNNNNNNNN | 1 |
| D | 589 hCoV-19/Spain/AR-IBV-2399/2020\|EPI_ISL_468802\|2020 | ···················· | ···················· | ···················· | ···················· | NNNNNNNNNNNNNNNNNNNN | NNNNNNNNNNNNNNNNNNNN | 1 |
| D | 590 hCoV-19/Spain/AR-IBV-2446/2020\|EPI_ISL_468803\|2020 | ···················· | ···················· | ···················· | ···················· | NNNNNNNNNNNNNNNNNNNN | NNNNNNNNNNNNNNNNNNNN | 1 |
| D | 591 hCoV-19/Spain/AR-IBV-2445/2020\|EPI_ISL_468808\|2020 | ···················· | ···················· | ···················· | ···················· | NNNNNNNNNNNNNNNNNNNN | NNNNNNNNNNNNNNNNNNNN | 1 |
| D | 592 hCoV-19/Spain/AR-IBV-2441/2020\|EPI_ISL_468809\|2020 | ···················· | ···················· | ···················· | ···················· | NNNNNNNNNNNNNNNNNNNN | NNNNNNNNNNNNNNNNNNNN | 1 |
| D | 593 hCoV-19/Spain/AR-IBV-2465/2020\|EPI_ISL_468810\|2020 | ···················· | ···················· | ···················· | ···················· | NNNNNNNNNNNNNNNNNNNN | NNNNNNNNNNNNNNNNNNNN | 1 |
| D | 594 hCoV-19/Spain/AR-IBV-2484/2020\|EPI_ISL_468811\|2020 | ···················· | ···················· | ···················· | ···················· | NNNNNNN············· | ···················· | 1 |
| D | 595 hCoV-19/Spain/AR-IBV-2438/2020\|EPI_ISL_468812\|2020 | ···················· | ···················· | ···················· | ···················· | NNNNNNNNNNNNNNNNNNNN | NNNNNNNNNNNNNNNN···· | 1 |
| D | 596 hCoV-19/Spain/AR-IBV-2479/2020\|EPI_ISL_468813\|2020 | ···················· | ···················· | ···················· | ···················· | NNNNNNNNNNNN········ | ···················· | 1 |
| D | 597 hCoV-19/Spain/AR-IBV-2464/2020\|EPI_ISL_468814\|2020 | ···················· | ···················· | ···················· | ···················· | ·NNNNNNNNNNNNNNNNNNN | NNNNNNNNNNNNNNNN···· | 1 |
| D | 599 hCoV-19/Spain/AR-IBV-2474/2020\|EPI_ISL_468816\|2020 | ···················· | ···················· | ···················· | ···················· | NNNNNNNNNNNNNNNNNNNN | NNNNNNNNNNNNNNNNNNNN | 1 |
| D | 600 hCoV-19/Spain/AR-IBV-2425/2020\|EPI_ISL_468817\|2020 | ···················· | ··········A········· | ···················· | ···················· | ··NNNNN············· | ···················· | 1 |
| D | 601 hCoV-19/Spain/AR-IBV-2437/2020\|EPI_ISL_468818\|2020 | ···················· | ···················· | ···················· | ···················· | NNNNNNN··A·········· | ···················· | 1 |
| D | 602 hCoV-19/Spain/AR-IBV-2444/2020\|EPI_ISL_468819\|2020 | ···················· | ···················· | ···················· | ···················· | NNNNNNNNNNNNNNNNN··· | ···················· | 1 |
| D | 603 hCoV-19/Spain/AR-IBV-2421/2020\|EPI_ISL_468820\|2020 | ···················· | ···················· | ···················· | ···················· | NNNNNNNNNNNNNNNNNNNN | NNNNNNNNNNNNNNNN···· | 1 |
| D | 604 hCoV-19/Spain/AR-IBV-2389/2020\|EPI_ISL_468822\|2020 | ···················· | ···················· | ···················· | ···················· | NNNNNNNNNNNNNNNNNNNN | NNNNNNNNNNNNNNNNNNNN | 1 |
| D | 605 hCoV-19/Spain/AR-IBV-2394/2020\|EPI_ISL_468823\|2020 | ···················· | ···················· | ···················· | ···················· | NNNNNNNNNNNNNNNNNNNN | NNNNNNNNNNNNNNNNNNNN | 1 |
| D | 606 hCoV-19/Spain/AR-IBV-2490/2020\|EPI_ISL_468824\|2020 | ···················· | ···················· | ···················· | ···················· | NNNNNNNNNNNNNNNNNNNN | NNNNNNNNNNNNNNNNNNNN | 1 |
| D | 607 hCoV-19/Spain/AR-IBV-2410/2020\|EPI_ISL_468825\|2020 | ···················· | ···················· | ···················· | ···················· | ··NNNNN············· | ·····NNNNNNNNNN····· | 1 |
| D | 608 hCoV-19/Spain/AR-IBV-2402/2020\|EPI_ISL_468826\|2020 | ···················· | ···················· | ···················· | ···················· | NNNNNNNNNNNNNNNNNNNN | NNNNNNNNNNNNNNNNNNNN | 1 |
| D | 609 hCoV-19/Spain/AR-IBV-2440/2020\|EPI_ISL_468828\|2020 | ···················· | ···················· | ···················· | ···················· | NNNNNNNNNNNNNNNNNNNN | NNNNNNNNNNNNNNNNNNNN | 1 |
| D | 610 hCoV-19/Spain/AR-IBV-2434/2020\|EPI_ISL_468829\|2020 | ···················· | ···················· | ···················· | ···················· | NNNNNNNNNNNNNNNNNNNN | NNNNNNNNNNNNNNNNNNNN | 1 |
| D | 611 hCoV-19/Spain/AR-IBV-2407/2020\|EPI_ISL_468830\|2020 | ···················· | ···················· | ···················· | ···················· | NNNNNNNNNNNNNNNNNNNN | NNNNNNNNNNNNNNNNNNNN | 1 |
| D | 613 hCoV-19/Spain/AR-IBV-2408/2020\|EPI_ISL_468832\|2020 | ···················· | ···················· | ···················· | ···················· | NNNNNNNNN··········· | ···················· | 1 |
| D | 614 hCoV-19/Spain/AR-IBV-2489/2020\|EPI_ISL_468833\|2020 | ···················· | ···················· | ···················· | ···················· | NNNNNNNNNNNNNNNNNNNN | NNNNNNNNNNNNNNNNNNNN | 1 |
| D | 615 hCoV-19/Spain/AR-IBV-2397/2020\|EPI_ISL_468834\|2020 | ···················· | ···················· | ···················· | ···················· | NNNNNNNNNNNNNNNNNNNN | NNNNNNNNNNNNNNNNNNNN | 1 |
| D | 616 hCoV-19/Spain/AR-IBV-2418/2020\|EPI_ISL_468835\|2020 | ···················· | ···················· | ···················· | ···················· | NNNNNNNNNNNNNNNNNNNN | NNNNNNNNNNNNNNNN·NNN | 1 |
| D | 617 hCoV-19/Spain/AR-IBV-2427/2020\|EPI_ISL_468836\|2020 | ···················· | ···················· | ···················· | ···················· | ··NNNNN············· | NNNNNNN······NN····· | 1 |
| D | 618 hCoV-19/Spain/AR-IBV-2385/2020\|EPI_ISL_468838\|2020 | ···················· | ···················· | ···················· | ···················· | NNNNNNNNNNNNNNNNNNNN | NNNNNNNNNNNNNNNNNNNN | 1 |
| D | 620 hCoV-19/Spain/AR-IBV-2426/2020\|EPI_ISL_468841\|2020 | ···················· | ···················· | ···················· | ···················· | NNNNNNNNNNNNNNNNNNNN | NNNNNNNNNNNNNNNNNNNN | 1 |
| D | 621 hCoV-19/Spain/AR-IBV-2419/2020\|EPI_ISL_468842\|2020 | ···················· | ···················· | ···················· | ···················· | NNNNNNNNNNNNNNNNNNNN | ···················· | 1 |
| D | 622 hCoV-19/Spain/AR-IBV-2430/2020\|EPI_ISL_468843\|2020 | ···················· | ···················· | ···················· | ···················· | NNNNNNNNNNNNNNNNNNNN | ···················· | 1 |
| D | 623 hCoV-19/Spain/AR-IBV-2413/2020\|EPI_ISL_468845\|2020 | ···················· | ···················· | ···················· | ···················· | NNNNNNNNNNNNNNNNNNNN | NNNNNNNNNNNNNNNNNNNN | 1 |
| D | 624 hCoV-19/Spain/AR-IBV-2463/2020\|EPI_ISL_468846\|2020 | ···················· | ···················· | ···················· | ···················· | NNNNNNNNNNNNNNNNNNNN | NNNNNNNNNNNNNNNNNNNN | 1 |
| D | 625 hCoV-19/Spain/AR-IBV-2473/2020\|EPI_ISL_468847\|2020 | ···················· | ···················· | ···················· | ···················· | NNNNNNNNNNNNNNNNNNNN | NNNNNNNNNNNNNNNNNNNN | 1 |
| D | 626 hCoV-19/Spain/AR-IBV-2439/2020\|EPI_ISL_468849\|2020 | ···················· | ···················· | ···················· | ···················· | NNNNNNNNNNNNNNNNNNNN | NNNNNNNNNNNNNNNNNNNN | 1 |
| D | 627 hCoV-19/Spain/AR-IBV-2477/2020\|EPI_ISL_468855\|2020 | ···················· | ···················· | ···················· | ···················· | NNNNNNNNNNNNNNNNNNNN | NNNNNNNNNNNNNNNNNNNN | 1 |
| D | 628 hCoV-19/Spain/AR-IBV-2447/2020\|EPI_ISL_468856\|2020 | ···················· | ···················· | ···················· | ···················· | NNNNNNNNNNNNNNNNNNNN | NNNNNNNNNNNNNNNNNNNN | 1 |
| D | 629 hCoV-19/Spain/AR-IBV-2412/2020\|EPI_ISL_468858\|2020 | ···················· | ···················· | ···················· | ···················· | NNNNNNN···NNNNNNNNNN | ···················· | 1 |
| D | 632 hCoV-19/Spain/PV-IBV-2117/2020\|EPI_ISL_468861\|2020 | ···················· | ···················· | ···················· | ···················· | ·NNNNNN············· | ···················· | 1 |
| D | 633 hCoV-19/Spain/PV-IBV-2162/2020\|EPI_ISL_468862\|2020 | ···················· | ···················· | ···················· | ···················· | NNNNNNNNNNNNNNNNNNNN | NNNNNNNNNNNNNNNNNNNN | 1 |
| D | 634 hCoV-19/Spain/PV-IBV-2100/2020\|EPI_ISL_468863\|2020 | ···················· | ···················· | ···················· | ···················· | NNNNNNNNNNNNNNNNNNNN | NNNNNNNNNNNNNNNNNNNN | 1 |
| D | 635 hCoV-19/Spain/PV-IBV-2074/2020\|EPI_ISL_468864\|2020 | ···················· | ···················· | ···················· | ···················· | NNNNNNNNNNNNNNNNNNNN | NNNNNNNNNNNNNNNNNNNN | 1 |
| D | 636 hCoV-19/Spain/PV-IBV-2141/2020\|EPI_ISL_468866\|2020 | ···················· | ···················· | ···················· | ···················· | NNNNNNNNNNNNNNNNNNNN | NNNNNNNNNNNNNNNNNNNN | 1 |

D 637 hCoV-19/Spain/PV-IBV-2164/2020|EPI_ISL_468867|2020 ···················· ···················· ···················· ···················· NNNNNNNNNNNNNNNNNNNN NNNNNNNNNNNNNNNNNNNN 1
D 638 hCoV-19/Spain/PV-IBV-2133/2020|EPI_ISL_468868|2020 ···················· ···················· ···················· ···················· NNNNNNNNNNNNNNNNNNNN ·····NNN······N····· 1
D 639 hCoV-19/Spain/PV-IBV-2116/2020|EPI_ISL_468869|2020 ···················· ···················· ···················· ···················· NNNNNNNNNNNNNNNNNNNN NNNNNNNNNNNNNNNNNNNN 1
D 640 hCoV-19/Spain/PV-IBV-2146/2020|EPI_ISL_468870|2020 ···················· ···················· ···················· ···················· NNNNNNNNNNNNNNNNNNNN NNNNNNNNNNNNNNNNNNNN 1
D 641 hCoV-19/Spain/PV-IBV-2160/2020|EPI_ISL_468871|2020 ···················· ···················· ···················· ···················· NNNNNNNNNNNNNNNNNNNN NNNNNNNNNNNNNNNNNNNN 1
D 642 hCoV-19/Spain/PV-IBV-2077/2020|EPI_ISL_468872|2020 ···················· ···················· ···················· ···················· NNNNNNNNNNNN········ ···················· 1
D 644 hCoV-19/Spain/PV-IBV-2110/2020|EPI_ISL_468874|2020 ···················· ···················· ···················· ···················· NNNNNNNNNNNNNNNNNNNN NNNNNNNNNNNNNNNNNNNN 1
D 645 hCoV-19/Spain/PV-IBV-2169/2020|EPI_ISL_468875|2020 ···················· ···················· ···················· ···················· NNNNNNNNNNNNNNNNNNNN NNNNNNNNNNNNNNNNNNNN 1
D 646 hCoV-19/Spain/PV-IBV-2085/2020|EPI_ISL_468876|2020 ···················· ···················· ···················· ···················· NNNNNNNNNNNNNNNNNNNN NNNNNNNNNNNNNNNNNNNN 1
D 647 hCoV-19/Spain/PV-IBV-2073/2020|EPI_ISL_468877|2020 ···················· ···················· ···················· ···················· NNNNNNNNNNNNNNNNNNNN NNNNNNNNNNNNNNNNNNNN 1
D 648 hCoV-19/Spain/PV-IBV-2130/2020|EPI_ISL_468878|2020 ···················· ···················· ···················· ···················· ·NNNNNNNNNNNNNNNNNNN NNNNNNNNNNNNNNN····· 1
D 649 hCoV-19/Spain/PV-IBV-2171/2020|EPI_ISL_468879|2020 ···················· ···················· ···················· ···················· NNNNNNNNNNNNNNNNNNNN NNNNNNNNNNNNNNNNNNNN 1
D 650 hCoV-19/Spain/PV-IBV-2111/2020|EPI_ISL_468880|2020 ···················· ···················· ···················· ···················· NNNNNNNNNNNNNNNNNNNN NNNNNNNNNNNNNNNNNNNN 1
D 652 hCoV-19/Spain/PV-IBV-2142/2020|EPI_ISL_468882|2020 ···················· ···················· ···················· ···················· NNNNNNNNNNNNNNNNNNNN ···················· 1
D 653 hCoV-19/Spain/PV-IBV-2101/2020|EPI_ISL_468883|2020 ···················· ···················· ···················· ···················· NNNNNNNNNNNNNNNNNNNN NNNNNNNNNNNNNNNNNNNN 1
D 655 hCoV-19/Spain/PV-IBV-2119/2020|EPI_ISL_468887|2020 ···················· ···················· ···················· ···················· ·NNNNNNNNNNNN······· ···················· 1
D 656 hCoV-19/Spain/PV-IBV-2093/2020|EPI_ISL_468888|2020 ···················· ···················· ···················· ···················· NNNNNNNNNNNNNNNNNNNN NNNNNNNNNNNNNNNNNNNN 1
D 657 hCoV-19/Spain/PV-IBV-2103/2020|EPI_ISL_468890|2020 ···················· ···················· ···················· ···················· NNNNNNNNNNNNNNNNNNNN ···················· 1
D 660 hCoV-19/Spain/PV-IBV-2158/2020|EPI_ISL_468893|2020 ···················· ···················· ···················· ···················· NNNNNNNNNNNNNNNNNNNN NNNNNNNNNNNNNNNNNNNN 1
D 661 hCoV-19/Spain/PV-IBV-2102/2020|EPI_ISL_468894|2020 ···················· ···················· ···················· ···················· NNNNNNNNNNNNNNNNNNNN NNNNNNNNNNNNNNNNNNNN 1
D 662 hCoV-19/Spain/PV-IBV-2152/2020|EPI_ISL_468895|2020 ···················· ···················· ···················· ···················· ·NNNNNN············· ···················· 1
D 663 hCoV-19/Spain/PV-IBV-2156/2020|EPI_ISL_468896|2020 ···················· ···················· ···················· ···················· NNNNNNNNNNNNNNNNNNNN NNNNNNNNNNNNNNNNNNNN 1
D 665 hCoV-19/Spain/PV-IBV-2132/2020|EPI_ISL_468898|2020 ···················· ···················· ···················· ···················· NNNNNNNNNNNNNNNNNNNN NNNNNNNNNNNNNNNNNNNN 1
D 688 hCoV-19/England/BIRM-5FB3E/2020|EPI_ISL_473377|202 ···················· ···················· ···················· ···················· NNNNNNNNNNNNNNNNNNNN NNNNNNNNNNNNNNNNNNNN 1
D 693 hCoV-19/England/BIRM-5FBA7/2020|EPI_ISL_473384|202 ···················· ···················· ···················· ···················· NNNNNNNNNNNNNNNNNNNN NNNNNNNNNNNNNNNNNNNN 1
D 707 hCoV-19/Singapore/454/2020|EPI_ISL_475985|2020-03- ···················· ···················· ···················· ···················· NNNNNNNNNNNNNNNNNNNN NNNNNNNNNNNNNNNNNNNN 1
D 733 hCoV-19/India/MH-NIV-18650/2020|EPI_ISL_479510|202 ···················· ···················· ···················· ·········NNNNNN····· ···················· ···················· 1
D 738 hCoV-19/India/MH-NIV-20878/2020|EPI_ISL_479515|202 ···················· ···················· ···················· ···········GA·NNNNNN ···················· ···················· 1
D 739 hCoV-19/India/MH-NIV-21041/2020|EPI_ISL_479516|202 ···················· ···················· ···················· ···NNNNNNNNNNNNNNNNN ···················· ···················· 1
D 747 hCoV-19/India/MH-NIV-25950/2020|EPI_ISL_479524|202 ···················· NNNNNNNN············ ···················· ···················· ···················· ···················· 1
D 748 hCoV-19/India/MH-NIV-27109/2020|EPI_ISL_479526|202 ···················· ···················· ···················· ·········NNNNNN····· ···················· ···················· 1
D 750 hCoV-19/India/MH-NIV-28416/2020|EPI_ISL_479528|202 ···················· ···················· ···················· ·········NNNNNNN···· ···················· ···················· 1
D 752 hCoV-19/India/MH-NIV-28900/2020|EPI_ISL_479530|202 ···················· ···················· ···················· ···TNNNNNNNNNNNNNNNN ···················· ···················· 1
D 754 hCoV-19/India/MH-NIV-29909/2020|EPI_ISL_479532|202 ···················· ···················· ···················· ·········NNNNNN····· ···················· ···················· 1
D 756 hCoV-19/India/MH-NIV-35868/2020|EPI_ISL_479534|202 ···················· ···················· ···················· ·········NNNNNNNNNNN ···················· ···················· 1
D 757 hCoV-19/India/MH-NIV-36405/2020|EPI_ISL_479535|202 ···················· ···················· ···················· ·········NNNNNN····· ···················· ···················· 1
D 764 hCoV-19/India/MH-NIV-38793/2020|EPI_ISL_479542|202 ···················· ···················· ···················· NNNNNNNNNNNNNNNNNNNN ···················· ···················· 1
D 769 hCoV-19/Canada/QC_AO3/2020|EPI_ISL_463920|2020-04- ···················· ···················· ···················· ···················· NNNNNNNNNNNNNNNNNNNN NNNNNNNNNNNNNNNNNNNN 1
D 773 hCoV-19/Canada/QC_AQ4/2020|EPI_ISL_463928|2020-04- ···················· NNNNNNNNNNNNNNNNNNNN ···················· ···················· ···················· ···················· 1
D 778 hCoV-19/Canada/QC_AR9/2020|EPI_ISL_463934|2020-04- ···················· NNNNNNNNNNNNNNNNNNNN ···················· ···················· ···················· ···················· 1
D 781 hCoV-19/Canada/QC_AT6/2020|EPI_ISL_463941|2020-04- ···················· NNNNNNNNNNNNNNNNNNNN ···················· ···················· ···················· ···················· 1
D 822 hCoV-19/England/20149009504/2020|EPI_ISL_466147|20 ···················· ···················· ···················· ···················· NNNNNNNNNNNNNNNNNNNN NNNNNNNNNNNNNNNNNNNN 1
D 823 hCoV-19/England/20149009604/2020|EPI_ISL_466148|20 ···················· ···················· ···················· ···················· ·NNNN······NNNNNN·NN NNNNN·N·N··N·····N·· 1
D 828 hCoV-19/England/20149012804/2020|EPI_ISL_466155|20 ···················· ···················· ···················· ···················· NNNNNNNNNNNNNNNNNNNN NNNNNNNNNNNNNNNNNNNN 1
D 852 hCoV-19/Spain/AR-IBV-2486/2020|EPI_ISL_468804|2020 ···················· ···················· ···················· ···················· NNNNNNNNNNNNNNNNNNNN NNNNNNNNNNNNNNN··NNN 1
D 853 hCoV-19/Spain/AR-IBV-2428/2020|EPI_ISL_468805|2020 ···················· ···················· ···················· ···················· NNNNNNN············· ···················· 1
D 854 hCoV-19/Spain/AR-IBV-2406/2020|EPI_ISL_468806|2020 ···················· ···················· ···················· ···················· NNNNNNNNNNNNNNNNNNNN NNNNNNNNNNNNNNNN···· 1
D 855 hCoV-19/Spain/AR-IBV-2459/2020|EPI_ISL_468807|2020 ···················· ···················· ···················· ···················· NNNNNNNNNNNNNNNNNNNN NNNNNNNNNNNNNNNNNNNN 1
D 898 hCoV-19/England/BIRM-5F8A0/2020|EPI_ISL_473345|202 ···················· ···················· ···················· ···················· NNNNNNNNNNNNNNNNNNNN NNNNNNNNNNNNNNNNNNNN 1
D 1029 hCoV-19/Canada/QC_AE4/2020|EPI_ISL_463905|2020-04- ···················· NNNNNNNNNNNNNNNNNNNN ···················· ···················· ···················· ···················· 1
D 1034 hCoV-19/Canada/QC_AF6/2020|EPI_ISL_463912|2020-04- ···················· NNNNNNNNNNNNNNNNNNNN ···················· ···················· ···················· ···················· 1
D 1035 hCoV-19/Canada/QC_AG4/2020|EPI_ISL_463913|2020-04- ···················· NNNNNNNNNNNNNNNNNNNN ···················· ···················· ···················· ···················· 1
D 1046 hCoV-19/England/20149008304/2020|EPI_ISL_466135|20 ···················· ···················· ···················· ···················· NNNNNNNNNNNNNNNNNNNN NNNNNNNNNNNNNNNNNNNN 1
D 1047 hCoV-19/England/20149008604/2020|EPI_ISL_466138|20 ···················· ···················· ···················· ···················· NNN·····NN·········· ············N······· 1
D 1052 hCoV-19/England/20149021804/2020|EPI_ISL_466164|20 ···················· ···················· ···················· ···················· ·NNNNNNNNNNNNNNNNNNN NNNNNNNNNNNNNNNNNNNN 1
D 1058 hCoV-19/England/20149023604/2020|EPI_ISL_466171|20 ···················· ···················· ···················· ···················· ···················· NNNN················ 1
D 1060 hCoV-19/England/20149024404/2020|EPI_ISL_466174|20 ···················· ···················· ···················· ···················· ·NNNNNNNNNNNNNNNNNNN NN·NN·····NNNNNNNNNN 1
D 1064 hCoV-19/England/20149028304/2020|EPI_ISL_466181|20 ···················· ···················· ···················· ···················· ···················· ·NNNN··············· 1
D 1065 hCoV-19/England/20149034604/2020|EPI_ISL_466185|20 ···················· ···················· ···················· ···················· NNNNNNNNNNNNNNNNNNNN N·N·····N·····NNNNNN 1
D 1067 hCoV-19/England/20149038204/2020|EPI_ISL_466189|20 ···················· ···················· ···················· ···················· NNNNNNNNNNNNNNNNNNNN ···················· 1
D 1069 hCoV-19/England/20149042204/2020|EPI_ISL_466192|20 ···················· ···················· ···················· ···················· NNNNNNNNNNNNNNNNNNNN NNNNNNNNNNNNNNNNNNNN 1
D 1072 hCoV-19/England/20149044204/2020|EPI_ISL_466197|20 ···················· ···················· ···················· ···················· NNNNNNNNNNNNNNNNNNNN NNNNNNNNNNNNNNNNNNNN 1
D 1073 hCoV-19/England/20149044304/2020|EPI_ISL_466198|20 ···················· ···················· ···················· ···················· ·NNNNNNNNNNNNNNNNNNN NNNNNNNNNNNNNNNNNNNN 1
D 1076 hCoV-19/Spain/MD-IBV-003671/2020|EPI_ISL_467201|20 ···················· ···················· ···················· ···················· NNNNNNNNNNNNNNNNNNNN NNNNNNNNNNNNNNNNNNNN 1
D 1077 hCoV-19/Spain/MD-IBV-003711/2020|EPI_ISL_467202|20 ···················· ···················· ···················· ···················· NNNNNNNNNNNNNNNNNNNN NNNNNNNNNNNNNNNNNNNN 1
D 1123 hCoV-19/Spain/AR-IBV-2436/2020|EPI_ISL_468778|2020 ···················· ···················· ···················· ···················· NNNNNNNNNNNNNNNNNNNN NNNNNNNNNNNNNNNNNNNN 1
D 1125 hCoV-19/Spain/AR-IBV-2485/2020|EPI_ISL_468780|2020 ···················· ···················· ···················· ···················· NNNNNNNNNNNNNNNNNNNN NNNNNNNNNNNNNNNNNNNN 1
D 1127 hCoV-19/Spain/AR-IBV-2409/2020|EPI_ISL_468782|2020 ···················· ···················· ···················· ···················· NNNNNNNNNNNN········ ···················· 1
D 1128 hCoV-19/Spain/AR-IBV-2450/2020|EPI_ISL_468783|2020 ···················· ···················· ···················· ···················· NNNNNNNNNNNNNNNNNNNN NNNNNNNNNNNNNNNNNNNN 1
D 1129 hCoV-19/Spain/AR-IBV-2471/2020|EPI_ISL_468784|2020 ···················· ···················· ···················· ···················· NNNNNNNNNNNNNNNNNNNN ···················· 1
D 1130 hCoV-19/Spain/AR-IBV-2483/2020|EPI_ISL_468785|2020 ···················· ···················· ···················· ···················· NNNNNNNNNNNNNNNNNNNN NNNNNNNNNNNNNNNNNNNN 1
D 1131 hCoV-19/Spain/AR-IBV-2388/2020|EPI_ISL_468786|2020 ···················· ···················· ···················· ···················· NNNNNNNNNNNNNNNNNNNN NNNNNNNNNNNNNNNNNNNN 1
D 1132 hCoV-19/Spain/AR-IBV-2432/2020|EPI_ISL_468788|2020 ···················· ···················· ···················· ···················· NNNNNNNNNNNNNNNNNNNN NNNNNNNNNNNNNNNNNNNN 1
D 1133 hCoV-19/Spain/AR-IBV-2469/2020|EPI_ISL_468790|2020 ···················· ···················· ···················· ···················· NNNNNNNNNNNNNNNNNNNN NNNNNNNNNNNNNNNNNNNN 1
D 1134 hCoV-19/Spain/AR-IBV-2458/2020|EPI_ISL_468791|2020 ···················· ···················· ···················· ···················· NNNNNNNNNNNNNNNNNNNN NNNNNNNNNNNNNNNNNNNN 1
D 1135 hCoV-19/Spain/AR-IBV-2455/2020|EPI_ISL_468792|2020 ···················· ···················· ···················· ···················· NNNNNNNNNNNNNNNNNNNN ···················· 1
D 1136 hCoV-19/Spain/AR-IBV-2429/2020|EPI_ISL_468793|2020 ···················· ···················· ···················· ···················· NNNNNNNNNNNNNNNNNNNN NNNNNNNNNNNNNNNNNNNN 1
D 1137 hCoV-19/Spain/AR-IBV-2478/2020|EPI_ISL_468794|2020 ···················· ···················· ···················· ···················· ·NNNNNN··NNNNNNNNNNN NNNNNNNNNNNNNNNNNNNN 1
D 1138 hCoV-19/Spain/AR-IBV-2398/2020|EPI_ISL_468795|2020 ···················· ···················· ···················· ···················· NNNNNNNNNNNNNNNNNNNN ···················· 1
D 1139 hCoV-19/Spain/AR-IBV-2417/2020|EPI_ISL_468797|2020 ···················· ···················· ···················· ···················· NNNNNNNNNNNNNNNNNNNN NNNNNNNNNNNNNNNNNNNN 1

| | | | | | | | | | |
|---|---|---|---|---|---|---|---|---|---|
| D | 1140 | hCoV-19/Spain/AR-IBV-2415/2020\|EPI_ISL_468798\|2020 | ···················· | ···········A········ | ···················· | ···················· | NNNNNNNNNNNNNNNNNNNN | ········NNNNNNNNNNNN | 1 |
| D | 1141 | hCoV-19/Spain/AR-IBV-2420/2020\|EPI_ISL_468799\|2020 | ···················· | ···················· | ···················· | ···················· | ·NNNNNNNNNNNNNNNNNNN | NNNNNNNNNNNNNNNNNNNN | 1 |
| D | 1207 | hCoV-19/Scotland/CVR3761/2020\|EPI_ISL_478105\|2020- | ···················· | ···················A | ···················· | ···················· | NNNNNNNNNNNNNNNNNNNN | ···················· | 1 |
| D | 1227 | hCoV-19/Scotland/CVR3807/2020\|EPI_ISL_478133\|2020- | ···················· | ···················· | ···················· | ···················· | NNNNNNNNNNNNNNNNNNNN | ···················· | 1 |
| D | 1279 | hCoV-19/India/MH-NIV-13307/2020\|EPI_ISL_479498\|202 | ···················· | ···················· | ···················· | ·········NNNNNNNNNNN | ···················· | ···················· | 1 |
| D | 1377 | hCoV-19/England/20118143104/2020\|EPI_ISL_464972\|20 | ···················· | ···················· | ···················· | ···················· | ·NNN····N··········· | ···················· | 1 |
| D | 1604 | hCoV-19/England/201180472/2020\|EPI_ISL_464917\|2020 | ···················· | ···················· | ···················· | ···················· | NNNNNNNNNNNNNNNNNNNN | NNNNNNNNNNNNNNNNNNNN | 1 |
| D | 1613 | hCoV-19/England/20118108504/2020\|EPI_ISL_464950\|20 | ···················· | ···················· | ···················· | ···················· | ··N·NNNNNNNNNNNNNNNN | NNNNNNNNNNNNNNNNNNNN | 1 |
| D | 1614 | hCoV-19/England/20118108604/2020\|EPI_ISL_464951\|20 | ···················· | ···················· | ···················· | ···················· | NNNNNNNNNNNNNNNNNNNN | NNNNNNNNNNNNNNNNNNNN | 1 |
| D | 1615 | hCoV-19/England/20118111104/2020\|EPI_ISL_464954\|20 | ···················· | ···················· | ···················· | ···················· | ···················· | NNNNNNNNNNNNNNNN···· | 1 |
| D | 1616 | hCoV-19/England/20118111804/2020\|EPI_ISL_464955\|20 | ···················· | ···················· | ···················· | ···················· | NNNNNNNNNNNNNNNNNNNN | NNNNNNNNNNNNNNNNNNNN | 1 |
| D | 1617 | hCoV-19/England/20118111904/2020\|EPI_ISL_464956\|20 | ···················· | ···················· | ···················· | ···················· | ·N·················· | NNNN················ | 1 |
| D | 1621 | hCoV-19/England/20118128204/2020\|EPI_ISL_464961\|20 | ···················· | ···················· | ···················· | ···················· | NNNNNNNNNNNNNNNNNNNN | NNNNNNNNNNNNNNNNNNNN | 1 |
| D | 1622 | hCoV-19/England/20118128304/2020\|EPI_ISL_464962\|20 | ···················· | ···················· | ···················· | ···················· | ·NNNNNNNNNNNNNNNNNNN | NNNNNNNNNNNNNNNNNNNN | 1 |
| D | 1638 | hCoV-19/Spain/PV-IBV-001548/2020\|EPI_ISL_467183\|20 | ···················· | ···················· | ···················· | ···················· | ·NNNNNN············· | ···················· | 1 |
| D | 1640 | hCoV-19/Spain/MD-IBV-003675/2020\|EPI_ISL_467185\|20 | ···················· | ···················· | ···················· | ···················· | NNNNNNNNNNNNNNNNNNNN | NNNNNNNNNNNNNNNNNNNN | 1 |
| D | 1645 | hCoV-19/Spain/MD-IBV-003703/2020\|EPI_ISL_467191\|20 | ···················· | ···················· | ···················· | ···················· | NNNNNNNNNNNNNNNNN··N | NNNNNNNNNNNNNNN····· | 1 |
| D | 1646 | hCoV-19/Spain/MD-IBV-003680/2020\|EPI_ISL_467192\|20 | ···················· | ···················· | ···················· | ···················· | NNNNNNNNNNNNNNNNNNNN | NNNNNNNNNNNNNNNNNNNN | 1 |
| D | 1650 | hCoV-19/Spain/MD-IBV-003650/2020\|EPI_ISL_467196\|20 | ···················· | ···················· | ···················· | ···················· | NNNNNNNNNNNNNNNNNNNN | NNNNNNNNNNNNNNNNNNNN | 1 |
| D | 1651 | hCoV-19/Spain/MD-IBV-003645/2020\|EPI_ISL_467197\|20 | ···················· | ···················· | ···················· | ···················· | NNNNNNNNNNNNNNNNNNNN | NNNNNNNNNNNNNNNNNNNN | 1 |
| D | 1806 | hCoV-19/England/20118070504/2020\|EPI_ISL_464938\|20 | ···················· | ···················· | ···················· | ···················· | ·NNNNNNNNNNNNNNNNNNN | NNNNNNNNNNNNNNNNNNNN | 1 |
| D | 1808 | hCoV-19/England/20118102304/2020\|EPI_ISL_464947\|20 | ···················· | ···················· | ···················· | ···················· | ···················· | NNNNNNNNN·N········· | 1 |
| D | 1818 | hCoV-19/Spain/PV-IBV-001534/2020\|EPI_ISL_467109\|20 | ···················· | ···················· | ···················· | ···················· | NNNNNNNNNNNN········ | ···················· | 1 |
| D | 1828 | hCoV-19/Spain/PV-IBV-001568/2020\|EPI_ISL_467121\|20 | ···················· | ···················· | ···················· | ···················· | NNNNNNNNNNNNNNNNNNNN | ···················· | 1 |
| D | 2089 | hCoV-19/Spain/PV-IBV-001603/2020\|EPI_ISL_467139\|20 | ···················· | ···················· | ···················· | ···················· | ···················· | NNNNNNNNNNNNNNN····· | 1 |
| D | 2091 | hCoV-19/Spain/PV-IBV-001570/2020\|EPI_ISL_467141\|20 | ···················· | ···················· | ···················· | ···················· | ·NNNNNN·····NNNNNNNN | ···················· | 1 |
| D | 2356 | hCoV-19/Spain/MD-IBV-003644/2020\|EPI_ISL_467205\|20 | ···················· | ···················· | ···················· | ···················· | NNNNNNNNNNNNNNNNNNNN | NNNNNNNNNNNNNNNNNNNN | 1 |
| D | 2359 | hCoV-19/Spain/MD-IBV-003659/2020\|EPI_ISL_467208\|20 | ···················· | ···················· | ···················· | ···················· | NNNNNNNNNNNNNNNNNNNN | NNNNNNNNNNNNNNNNNNNN | 1 |
| D | 2361 | hCoV-19/Spain/MD-IBV-003701/2020\|EPI_ISL_467211\|20 | ···················· | ···················· | ···················· | ···················· | NNNNNNNNNNNNNNNNNNNN | NNNNNNNNNNNNNNNNNNNN | 1 |
| D | 2362 | hCoV-19/Spain/MD-IBV-003713/2020\|EPI_ISL_467212\|20 | ···················· | ···················· | ···················· | ···················· | NNNNNNNNNNNNNNNNNNN· | ···················· | 1 |
| D | 2364 | hCoV-19/Spain/MD-IBV-003655/2020\|EPI_ISL_467214\|20 | ···················· | ···················· | ···················· | ···················· | NNNNNNNNNNNNNNNNNNNN | NNNNNNNNNNNNNNNNNNNN | 1 |
| D | 2367 | hCoV-19/Spain/MD-IBV-003678/2020\|EPI_ISL_467217\|20 | ···················· | ···················· | ···················· | ···················· | NNNNNNNNNNNNNNNNNNNN | NN·················· | 1 |
| D | 2368 | hCoV-19/Spain/MD-IBV-003670/2020\|EPI_ISL_467218\|20 | ···················· | ···················· | ···················· | ···················· | NNNNNNN············· | ···················· | 1 |
| D | 2370 | hCoV-19/Spain/MD-IBV-003712/2020\|EPI_ISL_467220\|20 | ···················· | ···················· | ···················· | ···················· | NNNNNNNNNNNNNNNNNNNN | NNNNNNNNNNNNNNNNNNNN | 1 |
| D | 2371 | hCoV-19/Spain/MD-IBV-003674/2020\|EPI_ISL_467221\|20 | ···················· | ···················· | ···················· | ···················· | NNNNNNNNNNNNNNNNNNNN | NNNNNNNNNNNNNNNNNNNN | 1 |
| D | 2373 | hCoV-19/Spain/MD-IBV-003673/2020\|EPI_ISL_467223\|20 | ···················· | ···················· | ···················· | ···················· | NNNNNNNNNNNNNNNNNNNN | NNNNNNNNNNNNNNNNNNNN | 1 |
| D | 2375 | hCoV-19/Spain/MD-IBV-003656/2020\|EPI_ISL_467225\|20 | ···················· | ···················· | ···················· | ···················· | NNNNNNNNNNNNNNNNNNNN | NNNNNNNNNNNNNNNNNNNN | 1 |
| D | 2376 | hCoV-19/Spain/MD-IBV-003649/2020\|EPI_ISL_467226\|20 | ···················· | ···················· | ···················· | ···················· | NNNNNNNNNNNNNNNNNNNN | NNNNNNNNNNNNNNN····· | 1 |
| D | 2378 | hCoV-19/Spain/MD-IBV-003648/2020\|EPI_ISL_467228\|20 | ···················· | ···················· | ···················· | ···················· | NNNNNNN············· | ···················· | 1 |
| D | 2381 | hCoV-19/Spain/MD-IBV-003699/2020\|EPI_ISL_467231\|20 | ···················· | ···················· | ···················· | ···················· | NNNNNNN············· | NNNNNNNNNNNNNNN····· | 1 |
| D | 2382 | hCoV-19/Spain/MD-IBV-003666/2020\|EPI_ISL_467232\|20 | ···················· | ···················· | ···················· | ···················· | NNNNNNNNNNNNNNNNNNNN | NNNNNNNNNNNNNNNNNNNN | 1 |
| D | 2383 | hCoV-19/Spain/MD-IBV-003682/2020\|EPI_ISL_467233\|20 | ···················· | ···················· | ···················· | ···················· | NNNNNNNNNNNNNNNNNNNN | NNNNNNNNNNNNNNNNNNNN | 1 |
| D | 2386 | hCoV-19/Spain/MD-IBV-003720/2020\|EPI_ISL_467236\|20 | ···················· | ···················· | ···················· | ···················· | NNNNNNNNNNNNNNNNNNNN | NNNNNNNNNNNNNNNNNNNN | 1 |
| D | 2387 | hCoV-19/Spain/MD-IBV-003679/2020\|EPI_ISL_467237\|20 | ···················· | ···················· | ···················· | ···················· | NNNNNNNNNNNNNNNNNNNN | NNNNNNNNNNNNNNN····· | 1 |
| D | 2388 | hCoV-19/Spain/MD-IBV-003716/2020\|EPI_ISL_467238\|20 | ···················· | ···················· | ···················· | ···················· | ·NNNNNN············· | ···················· | 1 |
| D | 2390 | hCoV-19/Spain/MD-IBV-003689/2020\|EPI_ISL_467240\|20 | ···················· | ···················· | ···················· | ···················· | NNNNNNNNNNNNNNNNNNNN | NNNNNNNNNNNNNNNNNNNN | 1 |
| D | 2392 | hCoV-19/Spain/MD-IBV-003718/2020\|EPI_ISL_467242\|20 | ···················· | ···················· | ···················· | ···················· | NNNNNNNNNNNNNNNNNNNN | NNNNNNNNNNNNNNNNNNNN | 1 |
| D | 2393 | hCoV-19/Spain/MD-IBV-003695/2020\|EPI_ISL_467243\|20 | ···················· | ···················· | ···················· | ···················· | NNNNNNNNNNNNNNNNNNNN | NNNNNNNNNNNNNNNNNNNN | 1 |
| D | 2394 | hCoV-19/Spain/MD-IBV-003652/2020\|EPI_ISL_467244\|20 | ···················· | ···················· | ···················· | ···················· | NNNNNNNNNNNNNNNNNNNN | NNNNNNNNNNNNNNNNNNNN | 1 |
| D | 2397 | hCoV-19/Spain/MD-IBV-003710/2020\|EPI_ISL_467248\|20 | ···················· | ···················· | ···················· | ···················· | NNNNNNNNNNNNNNNNNNNN | NNNNNNNNNNNNNNNNNNNN | 1 |
| D | 2398 | hCoV-19/Spain/GA-IBV-002737/2020\|EPI_ISL_467274\|20 | ···················· | ···················· | ···················· | ···················· | NNNNNNNNNNNNNNNNNNNN | NNNNNNNNNNNNNNNNNNNN | 1 |
| D | 2399 | hCoV-19/Spain/GA-IBV-002717/2020\|EPI_ISL_467275\|20 | ···················· | ···················· | ···················· | ···················· | NNNNNNNNNNNNNNNNNNNN | ·················NNN | 1 |
| D | 2400 | hCoV-19/Spain/GA-IBV-002753/2020\|EPI_ISL_467277\|20 | ···················· | ···················· | ···················· | ···················· | NNNNNNNNNNNNNNNNNNNN | NNNNNNNNNNNNNNNNNNNN | 1 |
| D | 2401 | hCoV-19/Spain/GA-IBV-002740/2020\|EPI_ISL_467280\|20 | ···················· | ···················· | ···················· | ···················· | NNNNNNNNNNNNNNNNNNNN | NNNNNNNNNNNNNNNNNNNN | 1 |
| D | 2402 | hCoV-19/Spain/GA-IBV-002731/2020\|EPI_ISL_467281\|20 | ···················· | ···················· | ···················· | ···················· | NNNNNNNNNNNNNNNNNNNN | NNNNNNNNNNNNNNNNNNNN | 1 |
| D | 2403 | hCoV-19/Spain/GA-IBV-002732/2020\|EPI_ISL_467283\|20 | ···················· | ···················· | ···················· | ···················· | NNNNNNNNNNNNNNNNNNNN | NNNNNNNNNNNNNNNNNNNN | 1 |
| D | 2404 | hCoV-19/Spain/GA-IBV-002755/2020\|EPI_ISL_467284\|20 | ···················· | ···················· | ···················· | ···················· | NNNNNNNNNNNNNNNNNNNN | NNNNNNNNNNNNNNNNNNNN | 1 |
| D | 2405 | hCoV-19/Spain/GA-IBV-002734/2020\|EPI_ISL_467285\|20 | ···················· | ···················· | ···················· | ···················· | NNNNNNNNNNNNNNNNNNNN | NNNNNNNNNNNNNNNNNNNN | 1 |
| D | 2406 | hCoV-19/Spain/GA-IBV-002754/2020\|EPI_ISL_467288\|20 | ···················· | ···················· | ···················· | ···················· | NNNNNNNNNNNNNNNNNNNN | NNNNNNNNNNNNNNNNNNNN | 1 |
| D | 2408 | hCoV-19/Spain/GA-IBV-002728/2020\|EPI_ISL_467290\|20 | ···················· | ···················· | ···················· | ···················· | NNNNNNNNNNNNNNNNNNNN | ···················· | 1 |
| D | 2409 | hCoV-19/Spain/GA-IBV-002738/2020\|EPI_ISL_467291\|20 | ···················· | ···················· | ···················· | ···················· | NNNNNNNNNNNNNNNNNNNN | ···········NNNN····· | 1 |
| D | 2410 | hCoV-19/Spain/GA-IBV-002752/2020\|EPI_ISL_467292\|20 | ···················· | ···················· | ···················· | ···················· | NNNNNNNNNNNNNNNNNNNN | NNNNNNNNNNNNNNNNNNNN | 1 |
| D | 2411 | hCoV-19/Spain/GA-IBV-002713/2020\|EPI_ISL_467293\|20 | ···················· | ···················· | ···················· | ···················· | NNNNNNNNNNNNNNNNNNNN | NNNNN··············· | 1 |
| D | 2412 | hCoV-19/Spain/GA-IBV-002715/2020\|EPI_ISL_467294\|20 | ···················· | ···················· | ···················· | ···················· | NNNNNNNNNNN········· | NNNNNNNNNNNNNNN····· | 1 |
| D | 2414 | hCoV-19/Spain/GA-IBV-002729/2020\|EPI_ISL_467296\|20 | ···················· | ···················· | ···················· | ···················· | NNNNNNNNNNNNNNNNNNNN | ·············NNNN··· | 1 |
| D | 2415 | hCoV-19/Spain/GA-IBV-002719/2020\|EPI_ISL_467297\|20 | ···················· | ···················· | ···················· | ···················· | ·NNNNNN············· | ···················· | 1 |
| D | 2521 | hCoV-19/Egypt/MASRI-10/2020\|EPI_ISL_475752\|2020-05 | ···················· | ···················· | ···················· | ·······ANNNNNNNNG··· | ···················· | ···················· | 1 |
| D | 2533 | hCoV-19/Austria/CeMM0118/2020\|EPI_ISL_475769\|2020- | ···················· | ···················· | ···················· | ···················· | ·······NNNNNNNNNNNNN | NNNNNNNNNNNNNNNNNNNN | 1 |
| D | 2535 | hCoV-19/Austria/CeMM0227/2020\|EPI_ISL_475771\|2020- | ···················· | ·····Y·············· | ···················· | ···················· | ·······NNNNNNNNNNNNN | NNNNNNNNNNNNNNNNNNNN | 1 |
| D | 2564 | hCoV-19/England/20169021004/2020\|EPI_ISL_465210\|20 | ···················· | ···················· | ···················· | ···················· | NNNNNN·N·NN·N··NN··N | ···N················ | 1 |
| D | 2575 | hCoV-19/England/20169037504/2020\|EPI_ISL_465245\|20 | ···················· | ···················· | ···················· | ···················· | ···················· | NNNNNNNNNNNNNNNNNNNN | 1 |
| D | 2578 | hCoV-19/England/20169042304/2020\|EPI_ISL_465254\|20 | ···················· | ···················· | ···················· | ···················· | ···················· | ··NNN··N············ | 1 |
| D | 2581 | hCoV-19/England/20170000404/2020\|EPI_ISL_465261\|20 | ···················· | ···················· | ···················· | ···················· | NNNNNNNNNNNNNNNNNNNN | ···················· | 1 |
| D | 2583 | hCoV-19/England/20170000904/2020\|EPI_ISL_465263\|20 | ···················· | ···················· | ···················· | ···················· | NNNNNNNNNNNNNNNNNNNN | NNNNNNNNNNNNNNNNNNNN | 1 |
| D | 2592 | hCoV-19/England/20166014404/2020\|EPI_ISL_466535\|20 | ···················· | ···················· | ···················· | ···················· | NNNNNNNNNNNNNNNNNNNN | NNNNNNNNNNNNNNNNN··· | 1 |
| D | 2600 | hCoV-19/England/20166056604/2020\|EPI_ISL_466554\|20 | ···················· | ···················· | ···················· | ···················· | NNNNNNNNNNNNNNNNNNNN | NNNNNNNNNNNNNNNNNNNN | 1 |
| D | 2603 | hCoV-19/England/20166060104/2020\|EPI_ISL_466558\|20 | ···················· | ···················· | ···················· | ···················· | NNNNNNNNNNNNNNNNNNNN | NNNNNNNNNNNNNNNNNN·· | 1 |
| D | 2608 | hCoV-19/England/20166064804/2020\|EPI_ISL_466564\|20 | ···················· | ···················· | ···················· | ···················· | NNNNNNNNNNNNNNNNNNNN | NNNNNNNNNNNNNNNNNNNN | 1 |
| D | 2614 | hCoV-19/England/20166069404/2020\|EPI_ISL_466575\|20 | ···················· | ···················· | ···················· | ···················· | NNNNNNNNNNNNNNNNNNNN | ···················· | 1 |
| D | 2617 | hCoV-19/England/20166071704/2020\|EPI_ISL_466583\|20 | ···················· | ···················· | ···················· | ···················· | NNNNNNNNNNNNNNNNNNNN | NNNNNNNNNNNNNNNNNNNN | 1 |

| | | | | | | | | | |
|---|---|---|---|---|---|---|---|---|---|
| D | 2618 | hCoV-19/England/20166071904/2020\|EPI_ISL_466584\|20 | ···················· | ···················· | ···················· | ···················· | NNNNNNNNNNNNNNNNNNNN | NNNNNNNNNNNNNNNNNNNN | 1 |
| D | 2619 | hCoV-19/England/20166072004/2020\|EPI_ISL_466585\|20 | ···················· | ···················· | ···················· | ···················· | ·NNNNNNNNNNNNNNNNNNN | ···················· | 1 |
| D | 2620 | hCoV-19/England/20166072904/2020\|EPI_ISL_466586\|20 | ···················· | ···················· | ···················· | ···················· | ··NNNNNNNNNNNNNNNNNN | ···················· | 1 |
| D | 2622 | hCoV-19/England/20168000804/2020\|EPI_ISL_466589\|20 | ···················· | ···················· | ···················· | ···················· | ·NNNNNNNNNNNNNNNNNNN | NNNNNNNNNNNNNNNNNNNN | 1 |
| D | 2623 | hCoV-19/England/20168000904/2020\|EPI_ISL_466590\|20 | ···················· | ···················· | ···················· | ···················· | ··NNNNNNNNNNNNNNNNNN | ···················· | 1 |
| D | 2633 | hCoV-19/Spain/MD-IBV-003658/2020\|EPI_ISL_467250\|20 | ···················· | ···················· | ···················· | ···················· | NNNNNNNNNNNNNNNNNNNN | NNNNNNNNNNNNNNNNNNNN | 1 |
| D | 2634 | hCoV-19/Spain/MD-IBV-003690/2020\|EPI_ISL_467251\|20 | ···················· | ···················· | ···················· | ···················· | NNNNNNNNNNNNNNNNNNNN | NNNNNNNNNNNNNNNNNNNN | 1 |
| D | 2635 | hCoV-19/Spain/MD-IBV-003692/2020\|EPI_ISL_467252\|20 | ···················· | ···················· | ···················· | ···················· | NNNNNNNNNNNNNNNNNNNN | NNNNNNNNNNNNNNNNNNNN | 1 |
| D | 2636 | hCoV-19/Spain/MD-IBV-003672/2020\|EPI_ISL_467253\|20 | ···················· | ··········T········· | ···················· | ···················· | NNNNNNNNNNNNNNNNNNNN | NNNNNNNNNNNNNNNNNNNN | 1 |
| D | 2644 | hCoV-19/Spain/GA-IBV-002749/2020\|EPI_ISL_467262\|20 | ···················· | ···················· | ···················· | ···················· | NNNNNNNNNNNNNNNNNNNN | NNNNNNNNNNNNNNNNNNNN | 1 |
| D | 2646 | hCoV-19/Spain/GA-IBV-002745/2020\|EPI_ISL_467265\|20 | ···················· | ···················· | ···················· | ···················· | NNNNNNNNNNNNNNNNNNNN | NNNNNNNNNNNNNNNNNNNN | 1 |
| D | 2647 | hCoV-19/Spain/GA-IBV-002726/2020\|EPI_ISL_467266\|20 | ···················· | ···················· | ···················· | ···················· | NNNNNNNNNNNNNNNNNNNN | NNNNNNNNNNNNNNNNNNNN | 1 |
| D | 2649 | hCoV-19/Spain/GA-IBV-002748/2020\|EPI_ISL_467269\|20 | ···················· | ···················· | ···················· | ···················· | NNNNNNNNNNNNNNNNNNNN | NNNNNNNNNNNNNNNNNNNN | 1 |
| D | 2650 | hCoV-19/Spain/GA-IBV-002716/2020\|EPI_ISL_467271\|20 | ···················· | ···················· | ···················· | ···················· | ·NNNNNNNNNNNNNNNNNNN | NNNNNNNNNNNNNNNNNNNN | 1 |
| D | 2651 | hCoV-19/Spain/GA-IBV-002730/2020\|EPI_ISL_467272\|20 | ···················· | ···················· | ···················· | ···················· | NNNNNNNNNNNNNNNNNNNN | NNNNNNNNNNNNNNNNNNNN | 1 |
| D | 2652 | hCoV-19/Spain/GA-IBV-002720/2020\|EPI_ISL_467273\|20 | ···················· | ···················· | ···················· | ···················· | NNNNNNNNNNNNNNNNNNNN | NNNNNNNNNNNNNNNNNNNN | 1 |
| D | 2653 | hCoV-19/Spain/GA-IBV-002727/2020\|EPI_ISL_467276\|20 | ···················· | ···················· | ···················· | ···················· | NNNNNNNNNNNNNNNNNNNN | NNNNNNNNNNNNNNNNNNNN | 1 |
| D | 2655 | hCoV-19/Spain/GA-IBV-002718/2020\|EPI_ISL_467279\|20 | ···················· | ···················· | ···················· | ···················· | ·NNNNNNNNNNNNNNNNNNN | ·············NN····· | 1 |
| D | 2745 | hCoV-19/England/NORW-ED18E/2020\|EPI_ISL_472354\|202 | ···················· | ···················· | ···················· | ···················· | NNNNNNNNNNNNNNNNNNNN | NNNNNNNNNNNNNNNNNNNN | 1 |
| D | 2746 | hCoV-19/England/NORW-ED24F/2020\|EPI_ISL_472356\|202 | ···················· | ···················· | ···················· | ···················· | NNNNNNNNNNNNNNNNNNNN | NNNNNNNNNNNNNNNNNNNN | 1 |
| D | 2749 | hCoV-19/Scotland/CVR3549/2020\|EPI_ISL_473684\|2020- | ···················· | ···················· | ···················· | ···················· | NNNNNNNNNNNNNNNNNNNN | ···················· | 1 |
| D | 2750 | hCoV-19/Scotland/CVR3551/2020\|EPI_ISL_473686\|2020- | ···················· | ·A·················· | ···················· | ···················· | NNNNNNNNNNNNNNNNNNNN | ···················· | 1 |
| D | 2846 | hCoV-19/England/20169005304/2020\|EPI_ISL_465181\|20 | ···················· | ···················· | ···················· | ···················· | NNNNNNNNNNNNNNNNNNNN | ·NNNN·····N···N·N··· | 1 |
| D | 2847 | hCoV-19/England/20169005404/2020\|EPI_ISL_465182\|20 | ···················· | ···················· | ···················· | ···················· | ·····N·············· | NNNNNNNNNNNNNNNNNNNN | 1 |
| D | 2848 | hCoV-19/England/20169005704/2020\|EPI_ISL_465183\|20 | ···················· | ···················· | ···················· | ···················· | NNNNNNNNNNNNNNNNNNNN | NNNNNNNNNNNNNNNNNNNN | 1 |
| D | 2894 | hCoV-19/England/NORW-EB135/2020\|EPI_ISL_472301\|202 | ···················· | ···················· | ···················· | ···················· | NNNNNNNNNNNNNNNNNNNN | ···················· | 1 |
| D | 2895 | hCoV-19/England/NORW-EB162/2020\|EPI_ISL_472302\|202 | ···················· | ···················· | ···················· | ···················· | NNNNNNNNNNNNNNNNNNNN | NNNNNNN············· | 1 |
| D | 2896 | hCoV-19/England/NORW-EB171/2020\|EPI_ISL_472303\|202 | ···················· | ···················· | ···················· | ···················· | NNNNNNNNNNNNNNNNNNNN | ···················· | 1 |
| D | 2899 | hCoV-19/England/NORW-EB2D8/2020\|EPI_ISL_472307\|202 | ···················· | ···················· | ···················· | ···················· | NNNNNNNNNNNNNNNNNNNN | ···················· | 1 |
| D | 2902 | hCoV-19/England/NORW-EB751/2020\|EPI_ISL_472313\|202 | ···················· | ···················· | ···················· | ···················· | NNNNNNNNNNNNNNNNNNNN | ···················· | 1 |
| D | 2903 | hCoV-19/England/NORW-EB760/2020\|EPI_ISL_472314\|202 | ···················· | ···················· | ···················· | ···················· | NNNNNNNNNNNNNNNNNNNN | NNNNNNNNNNNNNNNNNNNN | 1 |
| D | 2904 | hCoV-19/England/NORW-EB7AC/2020\|EPI_ISL_472316\|202 | ···················· | ···················· | ···················· | ···················· | NNNNNNNNNNNNNNNNNNNN | ···················· | 1 |
| D | 2907 | hCoV-19/England/NORW-EB86D/2020\|EPI_ISL_472322\|202 | ···················· | ···················· | ···················· | ···················· | NNNNNNNNNNNNNNNNNNNN | NNNNNNNNNNNNNNNNNNNN | 1 |
| D | 2909 | hCoV-19/England/NORW-EC046/2020\|EPI_ISL_472328\|202 | ···················· | ···················· | ···················· | ···················· | NNNN················ | ···················· | 1 |
| D | 2910 | hCoV-19/England/NORW-ECA48/2020\|EPI_ISL_472330\|202 | ···················· | ···················· | ···················· | ···················· | NNNNNNNNNNNNNNNNNNNN | NNNNNNNNNNNNNNNN···· | 1 |
| D | 2911 | hCoV-19/England/NORW-ECB18/2020\|EPI_ISL_472334\|202 | ···················· | ···················· | ···················· | ···················· | NNNNNNNNNNNNNNNNNNNN | ···················· | 1 |
| D | 2914 | hCoV-19/England/NORW-ECD21/2020\|EPI_ISL_472343\|202 | ···················· | ···················· | ···················· | ···················· | NNNN················ | ···················· | 1 |
| D | 2915 | hCoV-19/England/NORW-ECE2E/2020\|EPI_ISL_472347\|202 | ···················· | ···················· | ···················· | ···················· | NNNNNNNNNNNNNNNNNNNN | NNNNNNNNNNNNNNNNNNNN | 1 |
| D | 2916 | hCoV-19/England/NORW-ECEC4/2020\|EPI_ISL_472349\|202 | ···················· | ···················· | ···················· | ···················· | NNNNNNNNNNNNNNNNNNNN | NNNNNNNNNNNNNNNNNNNN | 1 |
| D | 2917 | hCoV-19/England/NORW-ED32E/2020\|EPI_ISL_472361\|202 | ···················· | ···················· | ···················· | ···················· | NNNNNNNNNNNNNNNNNNNN | NNNNNNNNNNNNNNNNNNNN | 1 |
| D | 2918 | hCoV-19/England/NORW-ED3F1/2020\|EPI_ISL_472363\|202 | ···················· | ···················· | ···················· | ···················· | NNNNNNNNNNNNNNNNNNNN | NNNNNNNNNNNNNNNNNNNN | 1 |
| D | 2919 | hCoV-19/England/NORW-ED42B/2020\|EPI_ISL_472365\|202 | ···················· | ···················· | ···················· | ···················· | NNNNNNNNNNNNNNNNNNNN | NNNNNNNNNNNNNNNNNNNN | 1 |
| D | 2920 | hCoV-19/England/NORW-ED449/2020\|EPI_ISL_472366\|202 | ···················· | ···················· | ···················· | ···················· | NNNNNNNNNNNNNNNNNNNN | NNNNNNNNNNNNNNNN···· | 1 |
| D | 2921 | hCoV-19/England/NORW-ED458/2020\|EPI_ISL_472367\|202 | ···················· | ···················· | ···················· | ···················· | NNNNNNNNNNNNNNNNNNNN | NNNNNNNNNNNNNNNNNNNN | 1 |
| D | 2922 | hCoV-19/England/NORW-ED476/2020\|EPI_ISL_472368\|202 | ···················· | ···················· | ···················· | ···················· | NNNNNNNNNNNNNNNNNNNN | NNNNNNNNNNNNNNNNNNNN | 1 |
| D | 2923 | hCoV-19/England/NORW-ED4D0/2020\|EPI_ISL_472371\|202 | ···················· | ···················A | ···················· | ···················· | NNNNNNNNNNNNNNNNNNNN | NNNNNNNNNNNNNNNNNNNN | 1 |
| D | 2924 | hCoV-19/England/NORW-ED5A0/2020\|EPI_ISL_472375\|202 | ···················· | ···················· | ···················· | ···················· | NNNNNNNNNNNNNNNNNNNN | NNNNNNNNNNNNNNNNNNNN | 1 |
| D | 2925 | hCoV-19/England/NORW-ED5EC/2020\|EPI_ISL_472376\|202 | ···················· | ···················· | ···················· | ···················· | NNNNNNNNNNNNNNNNNNNN | NNNNNNNNNNNNNNNNNNNN | 1 |
| D | 2926 | hCoV-19/England/NORW-ED77D/2020\|EPI_ISL_472381\|202 | ···················· | ···················· | ···················· | ···················· | NNNNNNNNNNNNNNNNNNNN | NNNNNNNNNNNNNNNNNNNN | 1 |
| D | 2973 | hCoV-19/Scotland/CVR3508/2020\|EPI_ISL_473648\|2020- | ···················· | ···················· | ···················· | ···················· | NNNNNNNNNNNNNNNNNNNN | NNNNNNNNNNNNNNNNNNNN | 1 |
| D | 2974 | hCoV-19/Scotland/CVR3509/2020\|EPI_ISL_473649\|2020- | ···················· | ···················A | ···················· | ···················· | NNNNNNNNNNNNNNNNNNNN | NNNNNNNNNNNNNNNNNNNN | 1 |
| D | 2975 | hCoV-19/Scotland/CVR3511/2020\|EPI_ISL_473651\|2020- | ···················· | ···················· | ···················· | ···················· | NNNNNNNNNNNNNNNNNNNN | NNNNNNNNNNNNNNNNNNNN | 1 |
| D | 2987 | hCoV-19/Scotland/CVR3534/2020\|EPI_ISL_473672\|2020- | ···················· | ···················A | ···················· | ···················· | NNNNNNNNNNNNNNNNNNNN | NNNNNNNNNNNNNNNNNNNN | 1 |
| D | 2988 | hCoV-19/Scotland/CVR3536/2020\|EPI_ISL_473673\|2020- | ···················· | ···················· | ···················· | ···················· | NNNNNNNNNNNNNNNNNNNN | ···················· | 1 |
| D | 2992 | hCoV-19/Scotland/CVR3545/2020\|EPI_ISL_473680\|2020- | ···················· | ···················· | ···················· | ···················· | NNNNNNNNNNNNNNNNNNNN | NNNNNNNNNNNNNNNNNNNN | 1 |
| D | 2996 | hCoV-19/Spain/AN-IBV-002932/2020\|EPI_ISL_474900\|20 | ···················· | ···················· | ···················· | ···················· | NNNNNNNNNNNNNNNNNNNN | NNNNNNNNNNNNNNNNNNNN | 1 |
| D | 2997 | hCoV-19/Spain/CM-IBV-003753/2020\|EPI_ISL_474901\|20 | ···················· | ···················· | ···················· | ···················· | NNNNNNNNNNNNNNNNNNNN | NNNNNNNNNNNNNNNNNNNN | 1 |
| D | 2998 | hCoV-19/Spain/CM-IBV-003761/2020\|EPI_ISL_474902\|20 | ···················· | ···················· | ···················· | ···················· | NNNNNNNNNNNNNNNNNNNN | NNNNNNNNNNNNNNNNNNNN | 1 |
| D | 2999 | hCoV-19/Spain/CM-IBV-003784/2020\|EPI_ISL_474903\|20 | ···················· | ···················· | ···················· | ···················· | NNNNNNNNNNNNNNNNNNNN | NNNNNNNNNNNNNNNNNNNN | 1 |
| D | 3000 | hCoV-19/Spain/CM-IBV-003845/2020\|EPI_ISL_474904\|20 | ···················· | ···················· | ···················· | ···················· | NNNNNNNNNNNNNNNNNNNN | NNNNNNNNNNNNNNNNNNNN | 1 |
| D | 3001 | hCoV-19/Spain/AN-IBV-002983/2020\|EPI_ISL_474908\|20 | ···················· | ···················· | ···················· | ···················· | NNNNNNNNNNNNNNNNNNNN | ···················· | 1 |
| D | 3002 | hCoV-19/Spain/AN-IBV-002906/2020\|EPI_ISL_474909\|20 | ···················· | ···················· | ···················· | ···················· | ·NNNNNNNNNNNNNNNN··· | ···················· | 1 |
| D | 3003 | hCoV-19/Spain/AN-IBV-002953/2020\|EPI_ISL_474930\|20 | ···················· | ···················· | ···················· | ···················· | NNNNNNNNNNNNNNNNNNNN | ··············N····· | 1 |
| D | 3009 | hCoV-19/Spain/CM-IBV-003780/2020\|EPI_ISL_474940\|20 | ···················· | ···················· | ···················· | ···················· | NNNNNNNNNNNNNNNNNNNN | NNNNNNNNNNNNNNNNNNNN | 1 |
| D | 3010 | hCoV-19/Spain/AN-IBV-002980/2020\|EPI_ISL_474943\|20 | ···················· | ···················· | ···················· | ···················· | NNNNNNNNNNNNNNNNNNNN | NNNNNNNNNNNNNNNNNNNN | 1 |
| D | 3011 | hCoV-19/Spain/AN-IBV-003002/2020\|EPI_ISL_474944\|20 | ···················· | ···················· | ···················· | ···················· | NNNNNNNNNNNNNNNNNNNN | ···················· | 1 |
| D | 3012 | hCoV-19/Spain/CM-IBV-003768/2020\|EPI_ISL_474946\|20 | ···················· | ···················· | ···················· | ···················· | NNNNNNNNNNNNNNNNNNNN | NNNNNNNNNN·········· | 1 |
| D | 3013 | hCoV-19/Spain/CM-IBV-003765/2020\|EPI_ISL_474947\|20 | ···················· | ···················· | ···················· | ···················· | NNNNNNNNNNNNNNNNNNNN | NNNNNNNNNNNNNNNNNNNN | 1 |
| D | 3014 | hCoV-19/Spain/AN-IBV-002914/2020\|EPI_ISL_474948\|20 | ···················· | ···················· | ···················· | ···················· | NNNNNNNNNNNNNNNNNNNN | NNNNNNNNNNNNNNNNNNNN | 1 |
| D | 3015 | hCoV-19/Spain/AN-IBV-003019/2020\|EPI_ISL_474949\|20 | ···················· | ···················· | ···················· | ···················· | NNNNNNNNNNNNNNNNNNNN | NNNNNNNNNNNNNNNN···· | 1 |
| D | 3017 | hCoV-19/Spain/CM-IBV-003789/2020\|EPI_ISL_474951\|20 | ···················· | ···················· | ···················· | ···················· | NNNNNNNNNNNNNNNNNNNN | NNNNNNNNNNNNNNNNNNNN | 1 |
| D | 3018 | hCoV-19/Spain/CM-IBV-003744/2020\|EPI_ISL_474952\|20 | ···················· | ···················· | ···················· | ···················· | NNNNNNNNNNNNNNNNNNNN | ···················· | 1 |
| D | 3019 | hCoV-19/Spain/CM-IBV-003746/2020\|EPI_ISL_474953\|20 | ···················· | ···················· | ···················· | ···················· | NNNNNNNNNNNNNNNNNNNN | NNNNNNNNNNNNNNNNNNNN | 1 |
| D | 3020 | hCoV-19/Spain/CM-IBV-003790/2020\|EPI_ISL_474954\|20 | ···················· | ···················· | ···················· | ···················· | NNNNNNNNNNNNNNNNNNNN | NNNNNNNNNNNNNNNNN··· | 1 |
| D | 3021 | hCoV-19/Spain/CM-IBV-003847/2020\|EPI_ISL_474955\|20 | ···················· | ···················· | ···················· | ···················· | NNNNNNNNNNNNNNNNNNNN | NNNNNNNNNNNNNNNNNNNN | 1 |
| D | 3022 | hCoV-19/Spain/CM-IBV-003887/2020\|EPI_ISL_474956\|20 | ···················· | ···················· | ···················· | ···················· | NNNNNNNNNNNNNNNNNNNN | NNNNNNNNNNNNNNNNNNNN | 1 |
| D | 3113 | hCoV-19/England/20169013704/2020\|EPI_ISL_465192\|20 | ···················· | ···················· | ···················· | ···················· | NNNNNNNNNNNNNNNNNNNN | NNNNNNNNNNNNNNNNNNNN | 1 |
| D | 3114 | hCoV-19/England/20169013804/2020\|EPI_ISL_465193\|20 | ···················· | ···················· | ···················· | ···················· | ··NNNNNNNNNNNNNNNNNN | NNNNNNNNNNNNNNNNNNNN | 1 |
| D | 3115 | hCoV-19/England/20169013904/2020\|EPI_ISL_465194\|20 | ···················· | ···················· | ···················· | ···················· | ··NNNNNNNNNNNNNNNNNN | NNNNNNNNNNNNNNNNNNNN | 1 |
| D | 3118 | hCoV-19/England/20169014704/2020\|EPI_ISL_465197\|20 | ···················· | ···················· | ···················· | ···················· | NNNNNNNNNNNNNNNNNNNN | NNNNNNNNNNNNNNNNNNNN | 1 |

| | | | | | | | | |
|---|---|---|---|---|---|---|---|---|
| D | 3119 hCoV-19/England/20169014804/2020\|EPI_ISL_465198\|20 | ···················· | ···················· | ···················· | ···················· | ·NNNNNNNNNNNNNNNNNNN | NNNNNNNNNNNNNNNNNNNN | 1 |
| D | 3122 hCoV-19/England/20170048904/2020\|EPI_ISL_465304\|20 | ···················· | ···················· | ···················· | ···················· | ··NNNNNN··NNN·····NN | NNNNNNNNNNNNNNNNNNNN | 1 |
| D | 3123 hCoV-19/England/20170049804/2020\|EPI_ISL_465307\|20 | ···················· | ···················· | ···················· | ···················· | NNNNNNNNNNNNNNNNNNNN | NNNNNNNNNNNNNNNNNNNN | 1 |
| D | 3131 hCoV-19/England/20172002504/2020\|EPI_ISL_465320\|20 | ···················· | ···················· | ···················· | ···················· | ····N··············· | N·NN·N······N······· | 1 |
| D | 3135 hCoV-19/England/20172039304/2020\|EPI_ISL_465339\|20 | ···················· | ···················· | ···················· | ···················· | ··NN················ | NNNN·NN·····N··NN··· | 1 |
| D | 3144 hCoV-19/England/20174004304/2020\|EPI_ISL_465384\|20 | ···················· | ···················· | ···················· | ···················· | ·NNNNNNNNNNNNNNNNNNN | ··NNN·····N····NN··· | 1 |
| D | 3146 hCoV-19/England/20174009204/2020\|EPI_ISL_465388\|20 | ···················· | ···················· | ···················· | ···················· | ··NNNNNNNNNNNNNNNNNN | NNNN···············N | 1 |
| D | 3165 hCoV-19/USA/NE-10969/2020\|EPI_ISL_466658\|2020-05-0 | ···················· | ···················· | ···················· | ···················· | NNNNNNNNNNNNNNNNNNNN | NNNNNNNNNNNNNNNNNNNN | 1 |
| D | 3167 hCoV-19/USA/NE-10865/2020\|EPI_ISL_466665\|2020-05-0 | ···················· | ···················· | ···················· | ···················· | NNNNNNNNNNNNNNNNNNNN | NNNNNNNNNNNNNNNNNNNN | 1 |
| D | 3173 hCoV-19/USA/NE-10965/2020\|EPI_ISL_466671\|2020-05-0 | ···················· | ···················· | ···················· | ···················· | NNNNNNNNNNNNNNNNNNNN | NNNNNNNNNNNNNNNNNNNN | 1 |
| D | 3174 hCoV-19/USA/NE-11120/2020\|EPI_ISL_466672\|2020-05-0 | ···················· | ···················· | ···················· | ···················· | NNNNNNNNNNNNNNNNNNNN | NNNNNNNNNNNNNNNNNNNN | 1 |
| D | 3207 hCoV-19/Spain/CN-IBV-002594/2020\|EPI_ISL_474911\|20 | ···················· | ···················· | ···················· | ···················· | ·NNNNNNNNNNNNNNNNNNN | ···················· | 1 |
| D | 3208 hCoV-19/Spain/CN-IBV-002596/2020\|EPI_ISL_474912\|20 | ···················· | ···················· | ···················· | ···················· | ·NNNNNNNNNNN·NNNNNNN | ···················· | 1 |
| D | 3209 hCoV-19/Spain/CN-IBV-002597/2020\|EPI_ISL_474913\|20 | ···················· | ···················· | ···················· | ···················· | NNNNNNNNNNNNNNNNNNNN | NNNNNNNNNNNNNNNNNNNN | 1 |
| D | 3210 hCoV-19/Spain/CN-IBV-002598/2020\|EPI_ISL_474914\|20 | ···················· | ···················· | ···················· | ···················· | NNNNNNNNNNNNNNNNNNNN | NNNNNNNNNNNNNNNNNNNN | 1 |
| D | 3212 hCoV-19/Spain/CN-IBV-002602/2020\|EPI_ISL_474916\|20 | ···················· | ···················· | ···················· | ···················· | NNNNNNNNNNNNNNNNNNNN | NNNNNNNNNNNNNNNNNNNN | 1 |
| D | 3213 hCoV-19/Spain/CN-IBV-002604/2020\|EPI_ISL_474917\|20 | ···················· | ···················· | ···················· | ···················· | NNNNNNNNNNNNNNNNNNNN | NNNNNNNNNNNNNNNNNNNN | 1 |
| D | 3214 hCoV-19/Spain/CM-IBV-003787/2020\|EPI_ISL_474919\|20 | ···················· | ···················· | ···················· | ···················· | NNNNNNNNNNNNNNNNNNNN | NNNNNNNNNNNNNNNNNNNN | 1 |
| D | 3215 hCoV-19/Spain/MD-IBV-002881/2020\|EPI_ISL_474920\|20 | ···················· | ···················A | ···················· | ···················· | NNNNNNN············· | ···················· | 1 |
| D | 3216 hCoV-19/Spain/CM-IBV-003785/2020\|EPI_ISL_474921\|20 | ···················· | ···················· | ···················· | ···················· | NNNNNNNNNNNNNNNNNNNN | NNNNNNNNNNNNNNNNNNNN | 1 |
| D | 3217 hCoV-19/Spain/AN-IBV-002836/2020\|EPI_ISL_474922\|20 | ···················· | ···················· | ···················· | ···················· | NNNNNNNNNNNNNNNNNNNN | NNNNNNNNNNNNNNNNNNNN | 1 |
| D | 3218 hCoV-19/Spain/AN-IBV-002841/2020\|EPI_ISL_474923\|20 | ···················· | ···················· | ···················· | ···················· | NNNNNNNNNNNNNNNNNNNN | NNNNNNNNNNNNNNNNNNNN | 1 |
| D | 3219 hCoV-19/Spain/AN-IBV-002860/2020\|EPI_ISL_474926\|20 | ···················· | ···················· | ···················· | ···················· | NNNNNNN············· | ···················· | 1 |
| D | 3220 hCoV-19/Spain/AN-IBV-002862/2020\|EPI_ISL_474927\|20 | ···················· | ···················· | ···················· | ···················· | NNNNNNNNNNNNNNNNNNNN | NNNNNNNNNNNNNNNNNNNN | 1 |
| D | 3221 hCoV-19/Spain/AN-IBV-002937/2020\|EPI_ISL_474928\|20 | ···················· | ···················· | ···················· | ···················· | NNNNNNNNNNNNNNNNNNNN | NNNNNNNNNNNNNNN··NNN | 1 |
| D | 3352 hCoV-19/England/20168005704/2020\|EPI_ISL_466600\|20 | ···················· | ···················· | ···················· | ···················· | ·NNNNNNNNNNNNNNNNNNN | NNNNNNNNNNNNNNNNNNNN | 1 |
| D | 3355 hCoV-19/England/20168027004/2020\|EPI_ISL_466604\|20 | ···················· | ···················· | ···················· | ···················· | ·NNNNNNNNNNNNNNNNNNN | NNNNNNNNNNNNNNNNNNNN | 1 |
| D | 3356 hCoV-19/England/20168031304/2020\|EPI_ISL_466605\|20 | ···················· | ···················· | ···················· | ···················· | ·NNNNNNNNNNNNNNNNNNN | ····N··············· | 1 |
| D | 3359 hCoV-19/England/20168033004/2020\|EPI_ISL_466610\|20 | ···················· | ···················· | ···················· | ···················· | NNNNNNNNNNNNNNNNNNNN | ···················· | 1 |
| D | 3366 hCoV-19/England/20168038204/2020\|EPI_ISL_466618\|20 | ···················· | ···················· | ···················· | ···················· | ·NNNNNNNNNNNNNNNNNNN | NNNNNNNNNNNNNNNNNNNN | 1 |
| D | 3368 hCoV-19/England/20168038604/2020\|EPI_ISL_466620\|20 | ···················· | ···················· | ···················· | ···················· | ·NNNNNNNNNNNNNNNNNNN | NNNNNNNNNNNNNNNNNNNN | 1 |
| D | 3371 hCoV-19/England/20169000604/2020\|EPI_ISL_466624\|20 | ···················· | ···················· | ···················· | ···················· | NNNNNNNNNNNNNNNNNNNN | NNNNNNNNNNNNNNNNNNNN | 1 |
| D | 3378 hCoV-19/USA/NE-11314/2020\|EPI_ISL_466677\|2020-05-0 | ···················· | ················T··· | ···················· | ···················· | NNNNNNNNNNNNNNNNNNNN | NNNNNNNNNNNNNNNNNNNN | 1 |
| D | 3528 hCoV-19/Scotland/CVR3612/2020\|EPI_ISL_473732\|2020- | ···················· | ···················A | ···················· | ···················· | NNNNNNNNNNNNNNNNNNNN | NNNNNNNNNNNNNNNNNNNN | 1 |
| D | 3535 hCoV-19/Scotland/CVR3674/2020\|EPI_ISL_473754\|2020- | ···················· | ···················· | ···················· | ···················· | NNNNNNNNNNNNNNNNNNNN | ···················· | 1 |
| D | 3539 hCoV-19/Scotland/CVR3711/2020\|EPI_ISL_473766\|2020- | ···················· | ·A·················· | ···················· | ···················· | NNNNNNNNNNNNNNNNNNNN | ···················· | 1 |
| D | 3587 hCoV-19/England/20119004304/2020\|EPI_ISL_465012\|20 | ···················· | ···················· | ···················· | ···················· | NNNNNNNNNNNNNNNNNNNN | NNNNNNNNNNNNNNNNNNNN | 1 |
| D | 3588 hCoV-19/England/20119005204/2020\|EPI_ISL_465013\|20 | ···················· | ···················· | ···················· | ···················· | ·NNNNNNNNNNNNNNNNNNN | ···················· | 1 |
| D | 3592 hCoV-19/England/20170030504/2020\|EPI_ISL_465272\|20 | ···················· | ···················· | ···················· | ···················· | NNNNNNNNNNNNNNNNNNNN | NNNNNNNNNNNNNNNNNNNN | 1 |
| D | 3599 hCoV-19/England/20170032604/2020\|EPI_ISL_465279\|20 | ···················· | ···················· | ···················· | ···················· | ··NNNNNNNNNNNNNNNNNN | NNNNNNNNNNNNNNNNNNNN | 1 |
| D | 3601 hCoV-19/England/20170033904/2020\|EPI_ISL_465281\|20 | ···················· | ···················· | ···················· | ···················· | NNNNNNNNNNNNNNNNNNNN | NNNNNNNNNNNNNNNNNNNN | 1 |
| D | 3602 hCoV-19/England/20170035204/2020\|EPI_ISL_465282\|20 | ···················· | ···················· | ···················· | ···················· | NNNNNNNNNNNNNNNNNNNN | NNNNNNNNNNNNNNNNNNNN | 1 |
| D | 3604 hCoV-19/England/20170036204/2020\|EPI_ISL_465285\|20 | ···················· | ···················· | ···················· | ···················· | NNNNNNNNNNNNNNNNNNNN | ·NN·NN··NNN···NNNNNN | 1 |
| D | 3605 hCoV-19/England/20170036504/2020\|EPI_ISL_465286\|20 | ···················· | ···················· | ···················· | ···················· | ···················· | ·NNN··N············· | 1 |
| D | 3612 hCoV-19/England/20156036804/2020\|EPI_ISL_466308\|20 | ···················· | ···················· | ···················· | ···················· | ···················· | NNNNNNNNNNNNNNNNNNNN | 1 |
| D | 3615 hCoV-19/England/20156044904/2020\|EPI_ISL_466320\|20 | ···················· | ···················· | ···················· | ···················· | ····N··NNNNNNNNNNNNN | NNNNNNNNNNNNNNNNNNNN | 1 |
| D | 3616 hCoV-19/England/20156046604/2020\|EPI_ISL_466322\|20 | ···················· | ···················· | ···················· | ···················· | NNNNNNNNNNNNNNNNNNNN | NNNNNNNNNNNNNNNNNNNN | 1 |
| D | 3617 hCoV-19/England/20156047304/2020\|EPI_ISL_466323\|20 | ···················· | ···················· | ···················· | ···················· | ·NNNNNNNNNNNNNNNNNNN | NNNN················ | 1 |
| D | 3621 hCoV-19/England/20156048004/2020\|EPI_ISL_466327\|20 | ···················· | ···················· | ···················· | ···················· | ·NNNNNNNNNNNNNNNNNNN | NNNNNNNNNNNNNNNNNNNN | 1 |
| D | 3625 hCoV-19/England/20156051204/2020\|EPI_ISL_466340\|20 | ···················· | ···················· | ···················· | ···················· | ···N················ | NNNNNNNNNNNNNNNNNNNN | 1 |
| D | 3651 hCoV-19/Spain/PV-IBV-2120/2020\|EPI_ISL_468932\|2020 | ···················· | ···················· | ···················· | ···················· | NNNNNNNNNNNNNNNNNNNN | NNNNNNNNNNNNNNNNNNNN | 1 |
| D | 3652 hCoV-19/Spain/PV-IBV-2096/2020\|EPI_ISL_468933\|2020 | ···················· | ···················· | ···················· | ···················· | NNNNNNNNNNNNNN······ | ···················· | 1 |
| D | 3654 hCoV-19/Spain/PV-IBV-2078/2020\|EPI_ISL_468935\|2020 | ···················· | ···················· | ···················· | ···················· | NNNNNNNNNNNNNNNNNNNN | NNNNNNNNNNNNNNNNNNNN | 1 |
| D | 3655 hCoV-19/Spain/PV-IBV-2094/2020\|EPI_ISL_468936\|2020 | ···················· | ···················· | ···················· | ···················· | NNNNNNNNNNNNNNNNNNNN | NNNNNNNNNNNNNNNNNNNN | 1 |
| D | 3656 hCoV-19/Spain/PV-IBV-2157/2020\|EPI_ISL_468937\|2020 | ···················· | ···················· | ···················· | ···················· | NNNNNNNNNNNNNNNNNNNN | NNNNNNNNNNNNNNNNNNNN | 1 |
| D | 3657 hCoV-19/Spain/PV-IBV-2131/2020\|EPI_ISL_468938\|2020 | ···················· | ···················· | ···················· | ···················· | NNNNNNNNNNNNNNNNNNNN | NNNNNNNNNNNNNNNNNNNN | 1 |
| D | 3658 hCoV-19/Spain/PV-IBV-2134/2020\|EPI_ISL_468939\|2020 | ···················· | ···················· | ···················· | ···················· | NNNNNNNNNNNNNNNNNNNN | NNNNNNNNNNNNNNNNNNNN | 1 |
| D | 3661 hCoV-19/Spain/PV-IBV-2154/2020\|EPI_ISL_468942\|2020 | ···················· | ···················· | ···················· | ···················· | ·NNNNNNNNNNNNNNNNNNN | NNNNNNNNNNNNNNNNNNNN | 1 |
| D | 3662 hCoV-19/Spain/PV-IBV-2083/2020\|EPI_ISL_468943\|2020 | ···················· | ···················· | ···················· | ···················· | NNNNNNNNNNNNNNNNNNNN | NNNNNNNNNNNNNNN····· | 1 |
| D | 3663 hCoV-19/Spain/PV-IBV-2106/2020\|EPI_ISL_468944\|2020 | ···················· | ···················· | ···················· | ···················· | ·NNNNNN············· | ···················· | 1 |
| D | 3665 hCoV-19/Spain/PV-IBV-2091/2020\|EPI_ISL_468946\|2020 | ···················· | ···················· | ···················· | ···················· | NNNNNNNNNNNNNNNNNNNN | NNNNNNNNNNNNNNNNNNNN | 1 |
| D | 3666 hCoV-19/Spain/PV-IBV-2124/2020\|EPI_ISL_468947\|2020 | ···················· | ···················· | ···················· | ···················· | NNNNNNNNNNNNNNNNNNNN | NNNNNNNNNNNNNNNNNNNN | 1 |
| D | 3669 hCoV-19/Spain/PV-IBV-2115/2020\|EPI_ISL_468950\|2020 | ···················· | ···················· | ···················· | ···················· | NNNNNNNNNNNNNNNNNNNN | NNNNNNNNNNNNNNNNNNNN | 1 |
| D | 3670 hCoV-19/Spain/PV-IBV-2167/2020\|EPI_ISL_468951\|2020 | ···················· | ···················· | ···················· | ···················· | NNNNNNNNNNNNNNNNNNNN | ··············N····· | 1 |
| D | 3671 hCoV-19/Spain/IB-IBV-2577/2020\|EPI_ISL_468952\|2020 | ···················· | ···················· | ···················· | ···················· | NNNNNNNNNNNNNNNNNNNN | NNNNNNNNNNNNNNNNNNNN | 1 |
| D | 3672 hCoV-19/Spain/IB-IBV-2521/2020\|EPI_ISL_468953\|2020 | ···················· | ···················· | ···················· | ···················· | NNNNNNNNNNNNNNNNNNNN | ·········NNNNNN····· | 1 |
| D | 3673 hCoV-19/Spain/IB-IBV-h2536/2020\|EPI_ISL_468954\|202 | ···················· | ···················· | ···················· | ···················· | NNNNNNNNNNNNNNNNNNNN | NNNNNNNNNNNNNNNNNNNN | 1 |
| D | 3674 hCoV-19/Spain/IB-IBV-2524/2020\|EPI_ISL_468959\|2020 | ···················· | ···················· | ···················· | ···················· | NNNNNNNNNNNNNNNNNNNN | NNNNNNNNNNNNNNNNNNNN | 1 |
| D | 3675 hCoV-19/Spain/IB-IBV-2553/2020\|EPI_ISL_468960\|2020 | ···················· | ···················· | ···················· | ···················· | NNNNNNNNNNNNNNNNNNNN | NNNNNNNNNNNNNNNNNNNN | 1 |
| D | 3676 hCoV-19/Spain/IB-IBV-2504/2020\|EPI_ISL_468961\|2020 | ···················· | ···················· | ···················· | ···················· | NNNNNNNNNNNNNNNNNNNN | NNNNNNNNNNNNNNNNNNNN | 1 |
| D | 3677 hCoV-19/Spain/IB-IBV-2505/2020\|EPI_ISL_468962\|2020 | ···················· | ···················· | ···················· | ···················· | NNNNNNNNNNNNNNNNNNNN | NNNNNNNNNNNNNNNNNNNN | 1 |
| D | 3679 hCoV-19/Spain/IB-IBV-2546/2020\|EPI_ISL_468968\|2020 | ···················· | ···················· | ···················· | ···················· | NNNNNNNNNNNNNNNNNNNN | NNNNNNNNNNNNNNNNNNNN | 1 |
| D | 3680 hCoV-19/Spain/IB-IBV-2500/2020\|EPI_ISL_468972\|2020 | ···················· | ···················· | ···················· | ···················· | NNNNNNNNNNNNNNNNNNNN | NNNNNNNNNNNNNNNNNNNN | 1 |
| D | 3681 hCoV-19/Spain/IB-IBV-2497/2020\|EPI_ISL_468974\|2020 | ···················· | ···················· | ···················· | ···················· | NNNNNNNNNNNNNNNNNNNN | NNNNNNNNNNNNNNNNNNNN | 1 |
| D | 3683 hCoV-19/Spain/IB-IBV-2506/2020\|EPI_ISL_468976\|2020 | ···················· | ···················· | ···················· | ···················· | ··NNNNN············· | NNNNN········NN····· | 1 |
| D | 3684 hCoV-19/Spain/IB-IBV-2537/2020\|EPI_ISL_468977\|2020 | ···················· | ···················· | ···················· | ···················· | NNNNNNNNNNNNNNNNNNNN | NNNNNNNNNNNNNNNNNNNN | 1 |
| D | 3686 hCoV-19/Spain/IB-IBV-2571/2020\|EPI_ISL_468980\|2020 | ···················· | ···················· | ···················· | ···················· | ·NNNNNNNNNNN·NNNNNNN | NNNNNNNNNNNNNNNNN··· | 1 |
| D | 3687 hCoV-19/Spain/IB-IBV-h2502/2020\|EPI_ISL_468982\|202 | ···················· | ···················· | ···················· | ···················· | NNNNNNNNNNNNNNNNNNNN | NNNNNNNNN····NN····· | 1 |
| D | 3688 hCoV-19/Spain/IB-IBV-2545/2020\|EPI_ISL_468983\|2020 | ···················· | ···················· | ···················· | ···················· | NNNNNNNNNNNNNNNNNNNN | NNNNNNNNNNNNNNNNNNNN | 1 |
| D | 3689 hCoV-19/Spain/IB-IBV-2587/2020\|EPI_ISL_468984\|2020 | ···················· | ···················· | ···················· | ···················· | NNNNNNNNNNNNNNNNNNNN | NNNNNNNNNNNNNNNNNNNN | 1 |

```
D 3690 hCoV-19/Spain/IB-IBV-2568/2020|EPI_ISL_468986|2020 ···················· ···················· ···················· ···················· NNNNNNNNNNNNNNNNNNNN NNNNNNNNNNNNNNNNNNNN 1
D 3691 hCoV-19/Spain/IB-IBV-2541/2020|EPI_ISL_468987|2020 ···················· ·····A·············· ···················· ···················· NNNNNNNNNNNNNNNNNNNN NNNNNNNNNNNNNNNNNNNN 1
D 3692 hCoV-19/Spain/IB-IBV-2514/2020|EPI_ISL_468988|2020 ···················· ···················· ···················· ···················· NNNNNNNNNNNNNNNNNNNN NNNNNNNNNNNNNNNNNNNN 1
D 3693 hCoV-19/Spain/IB-IBV-2496/2020|EPI_ISL_468989|2020 ···················· ···················· ···················· ···················· NNNNNNNNNNNNNNNNNNNN ···················· 1
D 3694 hCoV-19/Spain/IB-IBV-2543/2020|EPI_ISL_468990|2020 ···················· ···················· ···················· ···················· NNNNNNNNNNNNNNNNNNNN NNNNNNNNNNNNNNNNNNNN 1
D 3696 hCoV-19/Spain/IB-IBV-2538/2020|EPI_ISL_468992|2020 ···················· ···················· ···················· ···················· NNNNNNNNNNNNNNNNNNNN NNNNNNNNNNNNNNNNNNNN 1
D 3697 hCoV-19/Spain/IB-IBV-2544/2020|EPI_ISL_468993|2020 ···················· ···················· ···················· ···················· NNNNNNNNNNNNNNNNNNNN NNNNNNNNNNNNNNNNNNNN 1
D 3698 hCoV-19/Spain/IB-IBV-2562/2020|EPI_ISL_468997|2020 ···················· ···················· ···················· ···················· NNNNNNNNNNNNNNNNNNNN NNNNNNNNNNNNNNNNNNNN 1
D 3699 hCoV-19/Spain/IB-IBV-2554/2020|EPI_ISL_468998|2020 ···················· ···················· ···················· ···················· NNNNNNNNNNNNNNNNNNNN NNNNNNNNNNNNNNN··NNN 1
D 3700 hCoV-19/Spain/IB-IBV-h2551/2020|EPI_ISL_468999|202 ···················· ···················· ···················· ···················· ·NNNNNN············· NNNNNNNNNNNNNNNNNNNN 1
D 3708 hCoV-19/Scotland/CVR3568/2020|EPI_ISL_473702|2020- ···················· ···················· ···················· ···················· NNNNNNNNNNNNNNNNNNNN ···················· 1
D 3711 hCoV-19/Scotland/CVR3574/2020|EPI_ISL_473707|2020- ···················· ···················A ···················· ···················· NNNNNNNNNNNNNNNNNNNN NNNNNNNNNNNNNNNNNNNN 1
D 3713 hCoV-19/Scotland/CVR3580/2020|EPI_ISL_473712|2020- ···················· ···················· ···················· ···················· NNNNNNNNNNNNNNNNNNNN ···················· 1
D 3717 hCoV-19/Scotland/CVR3590/2020|EPI_ISL_473719|2020- ···················· ···················A ···················· ···················· NNNNNNNNNNNNNNNNNNNN NNNNNNNNNNNNNNNNNNNN 1
D 3885 hCoV-19/England/20152055004/2020|EPI_ISL_466237|20 ···················· ···················· ···················· ···················· ··NNNNNNNNNNNNNNNNNN ···················· 1
D 3886 hCoV-19/England/20152066504/2020|EPI_ISL_466238|20 ···················· ···················· ···················· ···················· ··NNNNNNNNNNNNNNNNNN NNNNNNNNNNNNNNNNNNNN 1
D 3887 hCoV-19/England/20152066604/2020|EPI_ISL_466239|20 ···················· ···················· ···················· ···················· NNNNNNNNNNNNNNNNNNNN NNNNNNNNNNNNNNNNNNNN 1
D 3889 hCoV-19/England/20152068004/2020|EPI_ISL_466241|20 ···················· ···················· ···················· ···················· ·NNNNNNNNNNNNNNNNNNN NNNNNNNNNNNNNNNNNNNN 1
D 3893 hCoV-19/England/20154003004/2020|EPI_ISL_466250|20 ···················· ···················· ···················· ···················· NNNNNNNNNNNNNNNNNNNN NNNNNNNNNNNNNNNNNNNN 1
D 3898 hCoV-19/England/20154019904/2020|EPI_ISL_466257|20 ···················· ···················· ···················· ···················· ··NNNNNNNNNNNNNNNNNN ···················· 1
D 3901 hCoV-19/England/20154074604/2020|EPI_ISL_466260|20 ···················· ···················· ···················· ···················· NNNN·NNNN··N········ ···················· 1
D 3903 hCoV-19/England/20154075604/2020|EPI_ISL_466262|20 ···················· ···················· ···················· ···················· NNNNNNNNNNNNNNNNNNNN NNNNNNNNNNNNNNNNNNNN 1
D 3907 hCoV-19/England/20154083704/2020|EPI_ISL_466273|20 ···················· ···················· ···················· ···················· NNNNNNNNNNNNNNNNNNNN NNNNNNNNNNNNNNNNNNNN 1
D 3921 hCoV-19/Spain/PV-IBV-2079/2020|EPI_ISL_468900|2020 ···················· ···················· ···················· ···················· NNNNNNNNNNNNNNNNNNNN NNNNNNNNNNNNNNNNNNNN 1
D 3922 hCoV-19/Spain/PV-IBV-2098/2020|EPI_ISL_468902|2020 ···················· ···················· ···················· ···················· NNNNNNNNNNNNNNNNNNNN NNNNNNNNNNNNNNNNNNNN 1
D 3923 hCoV-19/Spain/PV-IBV-2143/2020|EPI_ISL_468903|2020 ···················· ···················· ···················· ···················· NNNNNNNNNNNNNNNNNNNN NNNNNNNNNNNNNNNNNNNN 1
D 3925 hCoV-19/Spain/PV-IBV-2107/2020|EPI_ISL_468905|2020 ···················· ···················· ···················· ···················· NNNNNNNNNNNNNNNNNNNN NNNNNNNNNNNNNNN····· 1
D 3926 hCoV-19/Spain/PV-IBV-2140/2020|EPI_ISL_468906|2020 ···················· ···················· ···················· ···················· ·NNNNNNNNNNN····N··· ···················· 1
D 3927 hCoV-19/Spain/PV-IBV-2109/2020|EPI_ISL_468907|2020 ···················· ···················· ···················· ···················· ·NNNNNN······NNNNNNN ···················· 1
D 3928 hCoV-19/Spain/PV-IBV-2159/2020|EPI_ISL_468908|2020 ···················· ···················· ···················· ···················· NNNNNNNNNNNNNNNNNNNN NNNNNNNNNNNNNNNNNNNN 1
D 3929 hCoV-19/Spain/PV-IBV-2136/2020|EPI_ISL_468909|2020 ···················· ···················· ···················· ···················· NNNNNNNNNNNNNNNNNNNN NNNNNNNNNNNNNNN····· 1
D 3931 hCoV-19/Spain/PV-IBV-2088/2020|EPI_ISL_468911|2020 ···················· ···················· ···················· ···················· NNNNNNNNNNNNNNNNNNNN NNNNNNNNNNNNNNNNNNNN 1
D 3933 hCoV-19/Spain/PV-IBV-2149/2020|EPI_ISL_468913|2020 ···················· ···················· ···················· ···················· NNNNNNNNNNNNNNNNNNNN NNNNNNNNNNNNNNN····· 1
D 3935 hCoV-19/Spain/PV-IBV-2150/2020|EPI_ISL_468915|2020 ···················· ···················· ···················· ···················· NNNNNNNNNNNNNNNNNNNN NNNNNNNNNNNNNNNNNNNN 1
D 3936 hCoV-19/Spain/PV-IBV-2125/2020|EPI_ISL_468916|2020 ···················· ···················· ···················· ···················· NNNNNNNNNNNNNNNNNNNN NNNNNNNNNNNNNNNNNNNN 1
D 3938 hCoV-19/Spain/PV-IBV-2155/2020|EPI_ISL_468918|2020 ···················· ···················· ···K················ ···················· NNNNNNNNNNNNNNNNNNNN NNNNNNNNNNNNNNNNNNNN 1
D 3939 hCoV-19/Spain/PV-IBV-2095/2020|EPI_ISL_468919|2020 ···················· ···················· ···················· ···················· ·NNNNNN···NNNNNNN··· ···················· 1
D 3940 hCoV-19/Spain/PV-IBV-2081/2020|EPI_ISL_468920|2020 ···················· ···················· ···················· ···················· ·NNNNNNNNNNNNNNNNNNN ·········N·········· 1
D 3942 hCoV-19/Spain/PV-IBV-2099/2020|EPI_ISL_468922|2020 ···················· ···················· ···················· ···················· NNNNNNNNNNNNNNNNNNNN NNNNNNNNNNNNNNNNNNNN 1
D 3943 hCoV-19/Spain/PV-IBV-2080/2020|EPI_ISL_468923|2020 ···················· ···················· ···················· ···················· NNNNNNNNNNNNNNNNNNNN NNNNNNNNNNNNNNNNNNNN 1
D 3944 hCoV-19/Spain/PV-IBV-2076/2020|EPI_ISL_468924|2020 ···················· ···················· ···················· ···················· NNNNNNNNNNNNNNNNNNNN NNNNNNNNNNNNNNNNNNNN 1
D 3945 hCoV-19/Spain/PV-IBV-2082/2020|EPI_ISL_468925|2020 ···················· ···················· ···················· ···················· NNNNNNNNNNNNNNNNNNNN NNNNNNNNNNNNNNNNNNNN 1
D 3946 hCoV-19/Spain/PV-IBV-2128/2020|EPI_ISL_468926|2020 ···················· ···················· ···················· ···················· ·NNNNNNNNNNNNNNNNNNN ···················· 1
D 3948 hCoV-19/Spain/PV-IBV-2147/2020|EPI_ISL_468928|2020 ···················· ···················· ···················· ···················· NNNNNNNNNNNNNNNNNNNN NNNNNNNNNNNNNNNNNNNN 1
D 3949 hCoV-19/Spain/PV-IBV-2144/2020|EPI_ISL_468929|2020 ···················· ···················· ···················· ···················· ·NNNNNNNNNNNNNN····· NNNNNNNNNNNNNNN····· 1
D 3950 hCoV-19/Spain/PV-IBV-2148/2020|EPI_ISL_468930|2020 ···················· ···················· ···················· ···················· NNNNNNNNNNNNNNNNNNNN NNNNNNNNNNNNNNNNNNNN 1
D 3951 hCoV-19/Spain/PV-IBV-2165/2020|EPI_ISL_468931|2020 ···················· ···················· ···················· ···················· NNNNNNNNNNNNNNNNNNNN ·········NNNNNN····· 1
D 3972 hCoV-19/England/LIVE-A0749/2020|EPI_ISL_472131|202 ···················· ···················· ···················· ···················· ·······NNNNNNNNNNNNN ···················· 1
D 3977 hCoV-19/England/BIRM-60341/2020|EPI_ISL_473400|202 ···················· ···················A ···················· ···················· NNNNNNNNNNNNNNNNNNNN NNNNNNNNNNNNNNNNNNNN 1
D 3978 hCoV-19/England/BIRM-6038D/2020|EPI_ISL_473403|202 ···················· ···················A ···················· ···················· NNNNNNNNNNNNNNNNNNNN NNNNNNNNNNNNNNNNNNNN 1
D 3979 hCoV-19/England/BIRM-603E7/2020|EPI_ISL_473406|202 ···················· ···················· ···················· ···················· NNNNNNNNNNNNNNNNNNNN NNNNNNNNNNNNNNNNNNNN 1
D 3980 hCoV-19/England/BIRM-603F6/2020|EPI_ISL_473407|202 ···················· ···················· ···················· ···················· NNNNNNNNNNNNNNNNNNNN NNNNNNNNNNNNNNNNNNNN 1
D 3981 hCoV-19/England/BIRM-60411/2020|EPI_ISL_473408|202 ···················· ···················· ···················· ···················· NNNNNNNNNNNNNNNNNNNN NNNNNNNNNNNNNNNNNNNN 1
D 3987 hCoV-19/England/BIRM-604C6/2020|EPI_ISL_473418|202 ···················· ···················· ···················· ···················· NNNNNNNNNNNNNNNNNNNN NNNNNNNNNNNNNNNNNNNN 1
D 3991 hCoV-19/England/BIRM-60569/2020|EPI_ISL_473424|202 ···················· ···················· ···················· ···················· NNNNNNNNNNNNNNNNNNNN NNNNNNNNNNNNNNNNNNNN 1
D 4001 hCoV-19/England/BIRM-61216/2020|EPI_ISL_473444|202 ···················· ···················· ···················· ···················· NNNNNNNNNNNNNNNNNNNN NNNNNNNNNNNNNNNNNNNN 1
D 4004 hCoV-19/England/BIRM-61252/2020|EPI_ISL_473447|202 ···················· ···················· ···················· ···················· NNNNNNNNNNNNNNNNNNNN NNNNNNNNNNNNNNNNNNNN 1
D 4005 hCoV-19/England/BIRM-6128F/2020|EPI_ISL_473448|202 ···················· ···················· ···················· ···················· NNNNNNNNNNNNNNNNNNNN NNNNNNNNNNNNNNNNNNNN 1
D 4024 hCoV-19/England/CAMB-1B4A66/2020|EPI_ISL_473492|20 ···················· ···················· ···················· ···················· NNNNNNNNNNNNNNNNNNNN NNNNNNNNNNNNNNNNNNNN 1
D 4039 hCoV-19/Spain/AN-IBV-002927/2020|EPI_ISL_474797|20 ···················· ···················· ···················· ···················· NNNNNNNNNNNNNNNNNNNN NNNNNNNNNNNNNNNNNNNN 1
D 4040 hCoV-19/Spain/CM-IBV-003749/2020|EPI_ISL_474798|20 ···················· ···················· ···················· ···················· NNNNNNNNNNNNNNNNNNNN NNNNNNNNNNNNNNNNNNNN 1
D 4068 hCoV-19/India/MH-NIV-45936/2020|EPI_ISL_479548|202 ···················· ···················· ···················· ·········ANNNNNNG··· ···················· ···················· 1
D 4071 hCoV-19/India/MH-NIV-47109/2020|EPI_ISL_479552|202 ···················· ···················· ···················· ·······A·ANNNNNNNNN· ···················· ···················· 1
D 4072 hCoV-19/India/MH-NIV-47470/2020|EPI_ISL_479553|202 ···················· ···················· ···················· ·········NNNNNN····· ···················· ···················· 1
D 4099 hCoV-19/England/20119073604/2020|EPI_ISL_465101|20 ···················· ···················· ···················· ···················· NNNNNNNNNNNNNNNNNNNN NNNNNNNNNNNNNNNNNNNN 1
D 4106 hCoV-19/England/20119092004/2020|EPI_ISL_465130|20 ···················· ···················· ···················· ···················· NNNNNNNNNNNNNNNNNNNN ····N··············· 1
D 4107 hCoV-19/England/20119092204/2020|EPI_ISL_465131|20 ···················· ···················· ···················· ···················· NNNNNNNNNNNNNNNNNNNN NNNNNNNNNNNNNNNNNNNN 1
D 4108 hCoV-19/England/20119094404/2020|EPI_ISL_465135|20 ···················· ···················· ···················· ···················· NNNNNNNNNNNNNNNNNNNN NNNNN·NN···N···N···· 1
D 4112 hCoV-19/England/20156001304/2020|EPI_ISL_466280|20 ···················· ···················· ···R················ ···················· ···················· NNNNNNNNNNNNNNNNNNNN 1
D 4113 hCoV-19/England/20156001504/2020|EPI_ISL_466282|20 ···················· ···················· ···················· ···················· ·NNNNNNNNNN···NNNNN· ··N··N·NNNNNNNNNNNNN 1
D 4115 hCoV-19/England/20156003604/2020|EPI_ISL_466290|20 ···················· ···················· ···················· ···················· ·NNNNNNNNNNNNNNNNNNN NNNNNNNNNNNNNNNNNNNN 1
D 4116 hCoV-19/England/20156003704/2020|EPI_ISL_466291|20 ···················· ···················· ···················· ···················· ·NNNNNNNNNNNNNNNNNNN NNNNNNNNNNNNNNNNNNNN 1
D 4122 hCoV-19/England/20162009704/2020|EPI_ISL_466408|20 ···················· ···················· ···················· ···················· NNNNNNNNNNNNNNNNNNNN NNNNNNNNNNNNNNNNNNNN 1
D 4125 hCoV-19/England/20162037504/2020|EPI_ISL_466415|20 ···················· ···················· ···················· ···················· NNNNNNNNNNNNNNNNNNNN ··N················· 1
D 4135 hCoV-19/England/20164067004/2020|EPI_ISL_466463|20 ···················· ···················· ···················· ···················· ··NN·N······N···N··· NNNNNNNNNNNNNNNNNNNN 1
D 4168 hCoV-19/England/NORW-EA50D/2020|EPI_ISL_472291|202 ···················· ···················· ···················· ···················· NNNNNNNNNNNNNNNNNNNN NNNNNNNNNNNNNNNNNNNN 1
D 4172 hCoV-19/Scotland/CVR2785/2020|EPI_ISL_473565|2020- ···················· ·A·················· ···················· ···················· NNNNNNNNNNNNNNNNNNNN ···················· 1
D 4197 hCoV-19/Scotland/CVR2844/2020|EPI_ISL_473591|2020- ···················· ···················· ···················· ···················· NNNNNNNNNNNNNNNNNNNN ···················· 1
D 4200 hCoV-19/Scotland/CVR2849/2020|EPI_ISL_473594|2020- ···················· ···················· ···················· ···················· NNNNNNNNNNNNNNNNNNNN ···················· 1
D 4206 hCoV-19/Spain/GA-IBV-002922/2020|EPI_ISL_474854|20 ···················· ···················· ···················· ···················· NNNNNNNNNNNNNNNNNNNN NNNNNNNNNNNNNNNNNNNN 1
```

```
D    4207 hCoV-19/Spain/AN-IBV-002949/2020|EPI_ISL_474855|20    ····················  ····················  ····················  ····················  NNNNNNNNNNNNNNNNNNNN  NNNNNNNNNNNNNNNNNNNN    1
D    4208 hCoV-19/Spain/AN-IBV-002848/2020|EPI_ISL_474857|20    ····················  ····················  ····················  ····················  NNNNNNNNNNNNNNNNNNNN  NNNNNNNNNNNNNNNNNNNN    1
D    4209 hCoV-19/Spain/AN-IBV-003022/2020|EPI_ISL_474894|20    ····················  ····················  ····················  ····················  NNNNNNNNNNNNNNNNNNNN  NNNNNNNNNNNNNNNNNNNN    1
D    4211 hCoV-19/Spain/AN-IBV-002992/2020|EPI_ISL_474898|20    ····················  ····················  ····················  ····················  NNNNNNNNNNNNNNNNNNNN  ····················    1
D    4285 hCoV-19/India/MH-NIV-64904/2020|EPI_ISL_479558|202    ····················  ····················  ····················  ·········NNNNNN·····  ····················  ····················    1
D    4419 hCoV-19/England/20119015604/2020|EPI_ISL_465031|20    ····················  ····················  ····················  ····················  ·NNN················  NNNN················    1
D    4420 hCoV-19/England/20119019304/2020|EPI_ISL_465033|20    ····················  ····················  ····················  ····················  ····················  NNNNNNNNNNNNNNN·····    1
D    4422 hCoV-19/England/20119025604/2020|EPI_ISL_465035|20    ····················  ····················  ····················  ····················  ··NNNNNNNNNNNNNNNNNN  NNNNNNNNNNNNNNNNNNNN    1
D    4426 hCoV-19/England/20119030104/2020|EPI_ISL_465041|20    ····················  ····················  ····················  ····················  ········NN··········  NNNNNNN···NNNNNNNNNN    1
D    4427 hCoV-19/England/20119030804/2020|EPI_ISL_465042|20    ····················  ····················  ····················  ····················  ····················  ···········NNNN·····    1
D    4429 hCoV-19/England/20119031604/2020|EPI_ISL_465044|20    ····················  ····················  ····················  ····················  NNNNNNNNNNNNNNNNNNNN  NNNNNNNNNNNNNNNNNNNN    1
D    4430 hCoV-19/England/20119032004/2020|EPI_ISL_465045|20    ····················  ····················  ····················  ····················  NNNNNNNNNNNNNNNNNNNN  NNNNNNNNNNNNNNNNNNNN    1
D    4437 hCoV-19/England/20119040504/2020|EPI_ISL_465058|20    ····················  ····················  ····················  ····················  ····················  NNNNNN·····NNNN·····    1
D    4440 hCoV-19/England/20119043104/2020|EPI_ISL_465063|20    ····················  ····················  ····················  ····················  ····················  NNNN················    1
D    4441 hCoV-19/England/20119044204/2020|EPI_ISL_465064|20    ····················  ····················  ····················  ····················  NNNNNNNNNNNNNNNNNNNN  NNNNNNNNNNNNNNNNNNNN    1
D    4477 hCoV-19/Scotland/CVR2603/2020|EPI_ISL_473525|2020-    ····················  ····················  ····················  ····················  NNNNNNNNNNNNNNNNNNNN  ····················    1
D    4501 hCoV-19/Scotland/CVR2766/2020|EPI_ISL_473555|2020-    ····················  ····················  ····················  ····················  NNNNNNNNNNNNNNNNNNNN  ····················    1
D    4504 hCoV-19/Scotland/CVR2774/2020|EPI_ISL_473559|2020-    ····················  ···················A  ····················  ····················  NNNNNNNNNNNNNNNNNNNN  ····················    1
D    4508 hCoV-19/Spain/CM-IBV-003755/2020|EPI_ISL_474801|20    ····················  ····················  ····················  ····················  NNNNNNNNNNNNNNNNNNNN  NNNNNNNNNNNNNNNNNNNN    1
D    4509 hCoV-19/Spain/CM-IBV-003757/2020|EPI_ISL_474802|20    ····················  ····················  ····················  ····················  NNNNNNNNNNNNNNNNNNNN  NNNNNNNNNNNNNNNNNNNN    1
D    4511 hCoV-19/Spain/CM-IBV-003778/2020|EPI_ISL_474811|20    ····················  ····················  ····················  ····················  NNNNNNNNNNNNNNNNNNNN  NNNNNNNNNNNNNNNNNNNN    1
D    4512 hCoV-19/Spain/CM-IBV-003779/2020|EPI_ISL_474812|20    ····················  ····················  ····················  ····················  NNNNNNNNNNNNNNNNNNNN  NNNNNNNNNNNNNNNNNNNN    1
D    4513 hCoV-19/Spain/CM-IBV-003781/2020|EPI_ISL_474813|20    ····················  ····················  ····················  ····················  NNNNNNNNNNNNNNNNNNNN  NNNNNNNNNNNNNNNNNNNN    1
D    4514 hCoV-19/Spain/CM-IBV-003786/2020|EPI_ISL_474815|20    ····················  ····················  ····················  ····················  NNNNNNN·············  ····················    1
D    4515 hCoV-19/Spain/CM-IBV-003788/2020|EPI_ISL_474816|20    ····················  ····················  ····················  ····················  NNNNNNNNNNNNNNNNNNNN  NNNNNNNNNNNNNNNNNNNN    1
D    4517 hCoV-19/Spain/CM-IBV-003814/2020|EPI_ISL_474819|20    ····················  ····················  ····················  ····················  NNNNNNNNNNNNNNNNNNNN  NNNNNNNNNNNNNNNNNNNN    1
D    4518 hCoV-19/Spain/CM-IBV-003816/2020|EPI_ISL_474820|20    ····················  ····················  ····················  ····················  NNNNNNNNNNNNNNNNNNNN  NNNNNNNNNNNNNNNNNNNN    1
D    4519 hCoV-19/Spain/CM-IBV-003850/2020|EPI_ISL_474822|20    ····················  ····················  ····················  ····················  NNNNNNNNNNNNNNNNNNNN  NNNNNNNNNNNNNNNNNNNN    1
D    4520 hCoV-19/Spain/CM-IBV-003851/2020|EPI_ISL_474823|20    ····················  ····················  ····················  ····················  NNNNNNNNNNNNNNNNNNNN  NNNNNNNNNNNNNNNNNNNN    1
D    4521 hCoV-19/Spain/CM-IBV-003879/2020|EPI_ISL_474824|20    ····················  ····················  ····················  ····················  NNNNNNNNNNNNNNNNNNNN  NNNNNNNNNNNNNNNNNNNN    1
D    4522 hCoV-19/Spain/CM-IBV-003882/2020|EPI_ISL_474825|20    ····················  ····················  ····················  ····················  NNNNNNNNNNNNNNNNNNNN  NNNNNNNNNNNNNNNNNNNN    1
D    4523 hCoV-19/Spain/CM-IBV-003883/2020|EPI_ISL_474826|20    ····················  ····················  ····················  ····················  NNNNNNNNNNNNNNNNNNNN  ····················    1
D    4524 hCoV-19/Spain/CM-IBV-003884/2020|EPI_ISL_474827|20    ····················  ····················  ····················  ····················  NNNNNNNNNNNNNNNNNNNN  NNN·················    1
D    4525 hCoV-19/Spain/CM-IBV-003885/2020|EPI_ISL_474828|20    ····················  ····················  ····················  ····················  NNNNNNNNNNNNNNNNNNNN  ···········NNNNNNNNN    1
D    4526 hCoV-19/Spain/CM-IBV-003894/2020|EPI_ISL_474830|20    ····················  ····················  ····················  ····················  ·NNNNNN·············  ····················    1
D    4527 hCoV-19/Spain/CM-IBV-003903/2020|EPI_ISL_474831|20    ····················  ····················  ····················  ····················  NNNNNNNNNNNNNNNNNNNN  NNNNNNNNNNNNNNNNNNNN    1
D    4528 hCoV-19/Spain/AN-IBV-003009/2020|EPI_ISL_474832|20    ····················  ····················  ····················  ····················  NNNNNNNNNNNNNNNNNNNN  NNNNNNNNNNNNNNNNNNNN    1
D    4529 hCoV-19/Spain/AN-IBV-002890/2020|EPI_ISL_474833|20    ····················  ····················  ····················  ····················  ·NNNNNN·············  ····················    1
D    4530 hCoV-19/Spain/AN-IBV-002902/2020|EPI_ISL_474834|20    ····················  ····················  ····················  ····················  NNNNNNNNNNNNNNNNNNNN  ·········NNNNNN·····    1
D    4532 hCoV-19/Spain/CM-IBV-003750/2020|EPI_ISL_474837|20    ····················  ····················  ····················  ····················  NNNNNNNNNNNNNNNNNNNN  NNNNNNNNNNNNNNNNNNNN    1
D    4533 hCoV-19/Spain/CM-IBV-003769/2020|EPI_ISL_474838|20    ····················  ····················  ····················  ····················  NNNNNNNNNNNNNNNNNNNN  NNNNNNNNNNNNNNNNNNNN    1
D    4534 hCoV-19/Spain/AN-IBV-002842/2020|EPI_ISL_474841|20    ····················  ····················  ····················  ····················  NNNNNNNNNNNNNNNNNNNN  NNNNNNNNNNNNNNNNNNNN    1
D    4535 hCoV-19/Spain/AN-IBV-002852/2020|EPI_ISL_474842|20    ····················  ····················  ····················  ····················  ····················  NNNNNNNN············    1
D    4536 hCoV-19/Spain/AN-IBV-002863/2020|EPI_ISL_474843|20    ····················  ····················  ····················  ····················  NNNNNNNNNNNNNNNNNNNN  NNNNNNNNNNNNNNNNNNNN    1
D    4538 hCoV-19/Spain/CM-IBV-003759/2020|EPI_ISL_474847|20    ····················  ····················  ····················  ····················  NNNNNNNNNNNNNNNNNNNN  NNNNNNNNNNNNNNNNNNNN    1
D    4539 hCoV-19/Spain/AN-IBV-003030/2020|EPI_ISL_474850|20    ····················  ····················  ····················  ····················  NNNNNNNNNNNNNNNNNNNN  NNNNNNNNNNNNNNNNNNNN    1
D    4540 hCoV-19/Spain/AN-IBV-002912/2020|EPI_ISL_474851|20    ····················  ····················  ····················  ····················  NNNNNNN·············  ····················    1
D    4541 hCoV-19/Spain/CN-IBV-002876/2020|EPI_ISL_474852|20    ····················  ····················  ····················  ····················  NNNNNNNNNNNNNNNNNNNN  ····················    1
D    4542 hCoV-19/Spain/CM-IBV-003771/2020|EPI_ISL_474853|20    ····················  ····················  ····················  ····················  NNNNNNNNNNNNNNNNNNNN  NNNNNNNNNNNNNNNNNNNN    1
D    4543 hCoV-19/Spain/AN-IBV-002974/2020|EPI_ISL_474856|20    ····················  ····················  ····················  ····················  NNNNNNNNNNNNNNNNNNNN  NNNNNNNNNNNNNNN·····    1
D    4544 hCoV-19/Spain/AN-IBV-002867/2020|EPI_ISL_474859|20    ····················  ····················  ····················  ····················  NNNNNNN·············  ····················    1
D    4545 hCoV-19/Spain/AN-IBV-002870/2020|EPI_ISL_474860|20    ····················  ····················  ····················  ····················  NNNNNNNNNNNNNNNNNNNN  ····················    1
D    4547 hCoV-19/Spain/AN-IBV-002879/2020|EPI_ISL_474864|20    ····················  ····················  ····················  ····················  NNNNNNNNNNNN··NNNNNN  ····················    1
D    4548 hCoV-19/Spain/AN-IBV-002889/2020|EPI_ISL_474867|20    ····················  ····················  ····················  ····················  NNNNNNNNNNNNNNNNN···  ····················    1
D    4553 hCoV-19/Spain/AN-IBV-002926/2020|EPI_ISL_474872|20    ····················  ····················  ····················  ····················  NNNNNNNNNNNNNNNNNNNN  NNNNNNNNNNNNNNNNNNNN    1
D    4554 hCoV-19/Spain/AN-IBV-002929/2020|EPI_ISL_474873|20    ····················  ····················  ····················  ····················  NNNNNNNNNNNNNNNNNNNN  NNNNNNNNNNNNNNN·····    1
D    4556 hCoV-19/Spain/AN-IBV-002931/2020|EPI_ISL_474875|20    ····················  ····················  ····················  ····················  NNNNNNNNNNNNNNNNNNNN  NNNNNNNNNNNNNNNNNNNN    1
D    4557 hCoV-19/Spain/AN-IBV-002940/2020|EPI_ISL_474876|20    ····················  ····················  ····················  ····················  NNNNNNNNNNNNNNNNNNNN  NNNNNNNNNNNNNNNNNNNN    1
D    4558 hCoV-19/Spain/AN-IBV-002952/2020|EPI_ISL_474879|20    ····················  ····················  ····················  ····················  NNNNNNNNNNNNNNNNNNNN  NNNNNNNNNNNNNNNNNNNN    1
D    4560 hCoV-19/Spain/AN-IBV-002959/2020|EPI_ISL_474881|20    ····················  ····················  ····················  ····················  NNNNNNNNNNNNNNNNNNNN  NNNNNNNNNNNNNNNNNNNN    1
D    4561 hCoV-19/Spain/AN-IBV-002963/2020|EPI_ISL_474882|20    ····················  ····················  ····················  ····················  NNNNNNNNNNNNNNNNNNNN  NNNNNNNNNNNNNNNNNNNN    1
D    4562 hCoV-19/Spain/AN-IBV-002964/2020|EPI_ISL_474883|20    ····················  ····················  ····················  ····················  NNNNNNNNNNNNNNNNNNNN  NNNNNNNNNNNNNNNNNNNN    1
D    4563 hCoV-19/Spain/AN-IBV-002986/2020|EPI_ISL_474886|20    ····················  ····················  ····················  ····················  NNNNNNNNNNNNNNNNNNNN  NNNNNNNNNNNNNNNNNNNN    1
D    4564 hCoV-19/Spain/AN-IBV-002989/2020|EPI_ISL_474887|20    ····················  ····················  ····················  ····················  NNNNNNNNNNNNNNNNNNNN  NNNNNNNNNNNNNNNNNNNN    1
D    4566 hCoV-19/Spain/AN-IBV-003007/2020|EPI_ISL_474889|20    ····················  ····················  ····················  ····················  NNNNNNNNNNNNNNNNN···  ··············N·····    1
D    4568 hCoV-19/Spain/AN-IBV-003014/2020|EPI_ISL_474891|20    ····················  ····················  ····················  ····················  NNNNNNNNNNNNNNNNNNNN  NNNNNNNNNNNNNNNNNNNN    1
D    4655 hCoV-19/India/OR-ILSCV15937/2020|EPI_ISL_463041|20    ····················  ····················  ····················  NNNNNNNNNNNNNNNNNNNN  ····················  ····················    1
D    4657 hCoV-19/England/201040691/2020|EPI_ISL_464342|2020    ····················  ····················  ····················  ····················  ····················  NNNNNNNNNNNNNNNNNNNN    1
D    4661 hCoV-19/England/201041398/2020|EPI_ISL_464348|2020    ····················  ····················  ····················  ····················  NNNNNNNNNNNNNNNNNNNN  NNNNNNNNNNNNNNNNNNNN    1
D    4667 hCoV-19/England/201060010/2020|EPI_ISL_464359|2020    ····················  ····················  ····················  ····················  NNNNNNNNNNNNNNNNNNNN  NNNNNNNNNNNNNNNNNNNN    1
D    4674 hCoV-19/England/201060025/2020|EPI_ISL_464368|2020    ····················  ····················  ····················  ····················  ··NNNNNNNNNNNNNNNNNN  ····················    1
D    4676 hCoV-19/England/201060027/2020|EPI_ISL_464370|2020    ····················  ····················  ····················  ····················  ··NNNNNNNNNNNNNNNNNN  NNNNNNNNNN···NN····N    1
D    4677 hCoV-19/England/201060032/2020|EPI_ISL_464371|2020    ····················  ····················  ····················  ····················  NNNNNNNNNNNNNNNNNNNN  NNNNNNNNNNNNNNNNNNNN    1
D    4679 hCoV-19/England/201060037/2020|EPI_ISL_464374|2020    ····················  ····················  ····················  ····················  ·NNNNNNNNNNNNNNNNNNN  NNNNNNNNNNNNNNNNNNNN    1
D    4687 hCoV-19/England/20119048004/2020|EPI_ISL_465071|20    ····················  ····················  ····················  ····················  ····················  NNNN················    1
D    4692 hCoV-19/England/20119058604/2020|EPI_ISL_465081|20    ····················  ····················  ····················  ····················  ··N·NNNNNNNNNNNNNNNN  ····················    1
D    4693 hCoV-19/England/20119059204/2020|EPI_ISL_465082|20    ····················  ·A··················  ····················  ····················  NNNNNNNNNNNNNNNNNNNN  NNNNNNNNNNNNNNNNNNNN    1
D    4701 hCoV-19/England/20119067904/2020|EPI_ISL_465093|20    ····················  ····················  ····················  ····················  ·····N·N·NN··N·····N  ·········N···NNNNNNN    1
D    4702 hCoV-19/England/20119070004/2020|EPI_ISL_465096|20    ····················  ····················  ····················  ····················  ·NNNNNNNNNNNNNNNNNNN  NNNNNNNNNNNNNNN·····    1
D    4705 hCoV-19/England/20212019304/2020|EPI_ISL_465676|20    ····················  ····················  ····················  ····················  ·NNNNNNNNNNNNNNNNNNN  NNNNNNNNNNNNNNNNNNNN    1
```

| | | | | | | | | | |
|---|---|---|---|---|---|---|---|---|---|
| D | 4731 | hCoV-19/Spain/CM-IBV-003767/2020\|EPI_ISL_474804\|20 | ···················· | ···················· | ···················· | ···················· | NNNNNNNNNNNNNNNNNNNN | NNNNNNNNNNNNNNNNNNNN | 1 |
| D | 4732 | hCoV-19/Spain/CM-IBV-003770/2020\|EPI_ISL_474805\|20 | ···················· | ···················· | ···················· | ···················· | NNNNNNNNNNNNNNNNNNNN | NNNNNNNNNNNNNNNNNNNN | 1 |
| D | 4733 | hCoV-19/Spain/CM-IBV-003772/2020\|EPI_ISL_474806\|20 | ···················· | ···················· | ···················· | ···················· | NNNNNNNNNNNNNNNNNNNN | NNNNNNNNNNNNNNNNNNNN | 1 |
| D | 4734 | hCoV-19/Spain/CM-IBV-003774/2020\|EPI_ISL_474807\|20 | ···················· | ···················· | ···················· | ···················· | NNNNNNNNNNNNNNNNNNNN | NNNNNNNNNNNNNNNNNNNN | 1 |
| D | 4735 | hCoV-19/Spain/CM-IBV-003775/2020\|EPI_ISL_474808\|20 | ···················· | ···················· | ···················· | ···················· | NNNNNNNNNNNNNNNNNNNN | NNNNNNNNNNNNNNNNNNNN | 1 |
| D | 4736 | hCoV-19/Spain/CM-IBV-003776/2020\|EPI_ISL_474809\|20 | ···················· | ···················· | ···················· | ···················· | NNNNNNNNNNNNNNNNNNNN | NNNNNNNNNNNNNNNNNNNN | 1 |
| D | 4770 | hCoV-19/Belgium/UGent-101/2020\|EPI_ISL_475061\|2020 | ···················· | ···················· | ···················· | ···················· | NNNNNNNNNNNNNNNNNNNN | NNNNNNNNNNNNNNNNNNNN | 1 |
| D | 4875 | hCoV-19/India/OR-ILSCV18556/2020\|EPI_ISL_463063\|20 | ···················· | ···················· | ···················· | NNNNNNNNNNNNNNNNNNNN | ···················· | ···················· | 1 |
| D | 4890 | hCoV-19/England/201060042/2020\|EPI_ISL_464378\|2020 | ···················· | ···················· | ···················· | ···················· | NNNNNNNNNNNNNNNNNNNN | NNNNNNNNNNNNNNNNNNNN | 1 |
| D | 4896 | hCoV-19/England/201060063/2020\|EPI_ISL_464390\|2020 | ···················· | ···················· | ···················· | ···················· | NNNNNNNNNNNNNNNNNNNN | NNNNNNNNNNNNNNNNNNNN | 1 |
| D | 4897 | hCoV-19/England/201060066/2020\|EPI_ISL_464392\|2020 | ···················· | ···················· | ···················· | ···················· | NNNNNNNNNNNNNNNNNNNN | NNNNNNNNNNNNNNNNNNNN | 1 |
| D | 4898 | hCoV-19/England/201060067/2020\|EPI_ISL_464393\|2020 | ···················· | ···················· | ···················· | ···················· | NNNNNNNNNNNNNNNNNNNN | NNNNNNNNNNNNNNNNNNNN | 1 |
| D | 4900 | hCoV-19/England/201060143/2020\|EPI_ISL_464396\|2020 | ···················· | ···················· | ···················· | ···················· | ···············N···· | NNNNNNNNNNNNNNN····· | 1 |
| D | 4903 | hCoV-19/England/20192073101/2020\|EPI_ISL_465600\|20 | ···················· | ···················· | ···················· | ···················· | ··NNNNNNNNNNNNNNNNNN | NNNNN····NNNNNNNNNNN | 1 |
| D | 4906 | hCoV-19/England/20192094001/2020\|EPI_ISL_465603\|20 | ···················· | ···················· | ···················· | ···················· | NNNNNNNNNNNNNNNNNNNN | NNNNNNNNNNNNNNNNNNNN | 1 |
| D | 4908 | hCoV-19/England/20194008304/2020\|EPI_ISL_465607\|20 | ···················· | ···················· | ···················· | ···················· | ··NNNNNNNNNNNNNNN··· | ···················· | 1 |
| D | 4909 | hCoV-19/England/20194020701/2020\|EPI_ISL_465608\|20 | ···················· | ···A················ | ···················· | ···················· | ·NNNNNNNNNNNNNNNNNNN | NNNNNNNNNNNNNNNNNNNN | 1 |
| D | 4913 | hCoV-19/England/20194103201/2020\|EPI_ISL_465613\|20 | ···················· | ···················· | ···················· | ···················· | ···················· | ···············N·NNN | 1 |
| D | 4914 | hCoV-19/England/20194104301/2020\|EPI_ISL_465614\|20 | ···················· | ···················· | ···················· | ···················· | ···················· | NNNNN··············· | 1 |
| D | 4918 | hCoV-19/England/20194106801/2020\|EPI_ISL_465618\|20 | ···················· | ···················· | ···················· | ···················· | ···················· | NNNNNNNNNNNNNNNNNNNN | 1 |
| D | 4920 | hCoV-19/England/20206139504/2020\|EPI_ISL_465665\|20 | ···················· | ···················· | ···················· | ···················· | ·NNNNNNN·NNN········ | ··N················· | 1 |
| D | 4921 | hCoV-19/England/20206139704/2020\|EPI_ISL_465666\|20 | ···················· | ···················· | ···················· | ···················· | ·NNNNNNNNNNNNNNNNNNN | NNNNNNNNNNNNNNNNNNNN | 1 |
| D | 4934 | hCoV-19/Switzerland/TI-ETHZ-100003/2020\|EPI_ISL_46 | ···················· | ···················· | ···················· | ···················· | ······NNNNNNNNNNNNNN | NNNNNNNNNNNNNNNNNNNN | 1 |
| D | 4935 | hCoV-19/Switzerland/BE-ETHZ-100004/2020\|EPI_ISL_46 | ···················· | ···················· | ···················· | ···················· | ······NNNNNNNNNNNNNN | NNNNNNNNNNNNNNNNNNNN | 1 |
| D | 4936 | hCoV-19/Switzerland/BL-ETHZ-100005/2020\|EPI_ISL_46 | ···················· | ···················· | ···················· | ···················· | ······NNNNNNNNNNNNNN | NNNNNNNNNNNNNNNNNNNN | 1 |
| D | 4937 | hCoV-19/Switzerland/BS-ETHZ-100006/2020\|EPI_ISL_46 | ···················· | ···················· | ···················· | ···················· | ······NNNNNNNNNNNNNN | ···················· | 1 |
| D | 4939 | hCoV-19/Switzerland/GR-ETHZ-100008/2020\|EPI_ISL_46 | ···················· | ···················· | ···················· | ···················· | ······SNNNNNNNNNNNNN | ···················· | 1 |
| D | 4940 | hCoV-19/Switzerland/BL-ETHZ-100010/2020\|EPI_ISL_46 | ···················· | ···················· | ···················· | ···················· | ······NNNNNNNNNNNNNN | ···················· | 1 |
| D | 4943 | hCoV-19/Switzerland/GR-ETHZ-100013/2020\|EPI_ISL_46 | ···················· | ···················· | ···················· | ···················· | ······NNNNNNNNNNNNNN | NNNNNNNNNNNNNNNNNNNN | 1 |
| D | 4944 | hCoV-19/Switzerland/BS-ETHZ-100014/2020\|EPI_ISL_46 | ···················· | ···················R | ···················· | ···················· | ······NNNNNNNNNNNNNN | ···················· | 1 |
| D | 4946 | hCoV-19/Switzerland/BS-ETHZ-100017/2020\|EPI_ISL_46 | ···················· | ···················· | ···················· | ···················· | ······NNNNNNNNNNNNNN | NNNNNNNNNNNNNNNNNNNN | 1 |
| D | 4947 | hCoV-19/Switzerland/BS-ETHZ-100018/2020\|EPI_ISL_46 | ···················· | ···················· | ···················· | ···················· | ······NNNNNNNNNNNNNN | ···················· | 1 |
| D | 4949 | hCoV-19/Switzerland/BL-ETHZ-100021/2020\|EPI_ISL_46 | ···················· | ···················· | ···················· | ···················· | ······NNNNNNNNNNNNNN | NNNNNNNNNNNNNNNNNNNN | 1 |
| D | 4950 | hCoV-19/Switzerland/BS-ETHZ-100022/2020\|EPI_ISL_46 | ···················· | ···················· | ···················· | ···················· | ······NNNNNNNNNNNNNN | NNNNNNNNNNNNNNNNNNNN | 1 |
| D | 4951 | hCoV-19/Switzerland/BE-ETHZ-100023/2020\|EPI_ISL_46 | ···················· | ···················· | ···················· | ···················· | ······NNNNNNNNNNNNNN | NNNNNNNNNNNNNNNNNNNN | 1 |
| D | 4952 | hCoV-19/Switzerland/VS-ETHZ-100024/2020\|EPI_ISL_46 | ···················· | ···················· | ···················· | ···················· | ······NNNNNNNNNNNNNN | NNNNNNNNNNNNNNNNNNNN | 1 |
| D | 4956 | hCoV-19/Switzerland/TI-ETHZ-100074/2020\|EPI_ISL_46 | ···················· | ···················· | ···················· | ···················· | ······NNNNNNNNNNNNNN | NNNNNNNNNNNNNNNNNNNN | 1 |
| D | 4957 | hCoV-19/Switzerland/UR-ETHZ-130001/2020\|EPI_ISL_46 | ···················· | ···················· | ···················· | ···················· | ······NNNNNNNNNNNNNN | NNNNNNNNNNNNNNNNNNNN | 1 |
| D | 4958 | hCoV-19/Switzerland/VD-ETHZ-130003/2020\|EPI_ISL_46 | ···················· | ·A·················· | ···················· | ···················· | ······NNNNNNNNNNNNNN | ···················· | 1 |
| D | 4990 | hCoV-19/Belgium/UGent-103/2020\|EPI_ISL_475063\|2020 | ···················· | ·········A·········· | ···················· | ···················· | NNNNNNNNNNNNNNNNNNNN | NNNNNNNNNNNNNNNNNNNN | 1 |
| D | 4991 | hCoV-19/Belgium/UGent-104/2020\|EPI_ISL_475064\|2020 | ···················· | ···················· | ···················· | ···················· | NNNNNNNNNNNNNNNNNNNN | NNNNNNNNNNNNNNNNNNNN | 1 |
| D | 4992 | hCoV-19/Belgium/UGent-105/2020\|EPI_ISL_475065\|2020 | ···················· | ···················· | ···················· | ···················· | NNNNNNNNNNNNNNNNNNNN | NNNNNNNNNNNNNNNNNNNN | 1 |
| D | 4993 | hCoV-19/Belgium/UGent-106/2020\|EPI_ISL_475066\|2020 | ···················· | ···················· | ···················· | ···················· | NNNNNNNNNNNNNNNNNNNN | NNNNNNNNNNNNNNNNNNNN | 1 |
| D | 4995 | hCoV-19/Belgium/UGent-108/2020\|EPI_ISL_475068\|2020 | ···················· | ···················· | ···················· | ···················· | NNNNNNNNNNNNNNNNNNNN | NNNNNNNNNNNNNNNNNNNN | 1 |
| D | 5122 | hCoV-19/England/201040021/2020\|EPI_ISL_464302\|2020 | ···················· | ···················· | ···················· | ···················· | NNNNNNNNNNNNNNNNNNNN | NNNNNNNNNNNNNNNNNNNN | 1 |
| D | 5123 | hCoV-19/England/201040027/2020\|EPI_ISL_464305\|2020 | ···················· | ···················· | ···················· | ···················· | ··NNNN·NNNNNNNNNNNNN | ···················· | 1 |
| D | 5127 | hCoV-19/England/20104004204/2020\|EPI_ISL_464312\|20 | ···················· | ···················· | ···················· | ···················· | NNNNNNNNNNNNNNNNNNNN | NNNNNNNNNNNNNNNNNNNN | 1 |
| D | 5129 | hCoV-19/England/201040046/2020\|EPI_ISL_464315\|2020 | ···················· | ···················· | ···················· | ···················· | NNNNNNNNNNNNNNNNNNNN | ····N·N·NN····N·N··· | 1 |
| D | 5131 | hCoV-19/England/201040077/2020\|EPI_ISL_464321\|2020 | ···················· | ···················· | ···················· | ···················· | ···················· | N·NNN·NN···N········ | 1 |
| D | 5139 | hCoV-19/England/20204052004/2020\|EPI_ISL_465649\|20 | ···················· | ···················· | ···················· | ···················· | ····NNN·NN·NN·N·N··· | ···················· | 1 |
| D | 5155 | hCoV-19/Germany/BY-MVP-0269/2020\|EPI_ISL_466911\|20 | ···················· | ···················· | ···················· | ···················· | NNNNNNNNNNNNNNNNNNNN | NNNNNNNNNNNNNNNNNNNN | 1 |
| D | 5156 | hCoV-19/Germany/BY-MVP-0273/2020\|EPI_ISL_466913\|20 | ···················· | ···················· | ···················· | ···················· | NNNNNNNNNNNNNNNNNNNN | NNNNNNNNNNNNNNNNNNNN | 1 |
| D | 5157 | hCoV-19/Switzerland/BS-ETHZ-100028/2020\|EPI_ISL_46 | ···················· | ···················· | ···················· | ···················· | ······NNNNNNNNNNNNNN | ·········Y·········· | 1 |
| D | 5159 | hCoV-19/Switzerland/BS-ETHZ-100030/2020\|EPI_ISL_46 | ···················· | ···················· | ···················· | ···················· | ······NNNNNNNNNNNNNN | NNNNNNNNNNNNNNNNNNNN | 1 |
| D | 5161 | hCoV-19/Switzerland/ZH-ETHZ-100032/2020\|EPI_ISL_46 | ···················· | ···················· | ···················· | ···················· | ······NNNNNNNNNNNNNN | ···················· | 1 |
| D | 5162 | hCoV-19/Switzerland/BS-ETHZ-100034/2020\|EPI_ISL_46 | ···················· | ···················· | ···················· | ···················· | ······NNNNNNNNNNNNNN | NNNNNNNNNNNNNNNNNNNN | 1 |
| D | 5166 | hCoV-19/Switzerland/TI-ETHZ-100039/2020\|EPI_ISL_46 | ···················· | ···················· | ···················· | ···················· | ······NNNNNNNNNNNNNN | NNNNNNNNNNNNNNNNNNNN | 1 |
| D | 5167 | hCoV-19/Switzerland/ZH-ETHZ-100041/2020\|EPI_ISL_46 | ···················· | ···················· | ···················· | ···················· | ······NNNNNNNNNNNNNN | NNNNNNNNNNNNNNNNNNNN | 1 |
| D | 5168 | hCoV-19/Switzerland/TI-ETHZ-100042/2020\|EPI_ISL_46 | ···················· | ···················· | ···················· | ···················· | ······NNNNNNNNNNNNNN | NNNNNNNNNNNNNNNNNNNN | 1 |
| D | 5169 | hCoV-19/Switzerland/TI-ETHZ-100043/2020\|EPI_ISL_46 | ···················· | ···················· | ···················· | ···················· | ······NNNNNNNNNNNNNN | NNNNNNNNNNNNNNNNNNNN | 1 |
| D | 5170 | hCoV-19/Switzerland/GR-ETHZ-100044/2020\|EPI_ISL_46 | ···················· | ···················· | ···················· | ···················· | ······NNNNNNNNNNNNNN | NNNNNNNNNNNNNNNNNNNN | 1 |
| D | 5171 | hCoV-19/Switzerland/TI-ETHZ-100045/2020\|EPI_ISL_46 | ···················· | ···················· | ···················· | ···················· | ······NNNNNNNNNNNNNN | NNNNNNNNNNNNNNNNNNNN | 1 |
| D | 5172 | hCoV-19/Switzerland/BE-ETHZ-100047/2020\|EPI_ISL_46 | ···················· | ···················· | ···················· | ···················· | ······NNNNNNNNNNNNNN | NNNNNNNNNNNNNNNNNNNN | 1 |
| D | 5174 | hCoV-19/Switzerland/TI-ETHZ-100049/2020\|EPI_ISL_46 | ···················· | ···················· | ···················· | ···················· | ······NNNNNNNNNNNNNN | NNNNNNNNNNNNNNNNNNNN | 1 |
| D | 5175 | hCoV-19/Switzerland/TI-ETHZ-100050/2020\|EPI_ISL_46 | ···················· | ···················· | ···················· | ···················· | ······NNNNNNNNNNNNNN | NNNNNNNNNNNNNNNNNNNN | 1 |
| D | 5176 | hCoV-19/Switzerland/TI-ETHZ-100051/2020\|EPI_ISL_46 | ···················· | ···················· | ···················· | ···················· | ······NNNNNNNNNNNNNN | ···················· | 1 |
| D | 5177 | hCoV-19/Switzerland/AG-ETHZ-100052/2020\|EPI_ISL_46 | ···················· | ···················· | ···················· | ···················· | ······NNNNNNNNNNNNNN | NNNNNNNNNNNNNNNNNNNN | 1 |
| D | 5178 | hCoV-19/Switzerland/BE-ETHZ-100053/2020\|EPI_ISL_46 | ···················· | ···················· | ···················· | ···················· | ······NNNNNNNNNNNNNN | NNNNNNNNNNNNNNNNNNNN | 1 |
| D | 5179 | hCoV-19/Switzerland/BE-ETHZ-100054/2020\|EPI_ISL_46 | ···················· | ···················· | ···················· | ···················· | ······NNNNNNNNNNNNNN | NNNNNNNNNNNNNNNNNNNN | 1 |
| D | 5180 | hCoV-19/Switzerland/BE-ETHZ-100055/2020\|EPI_ISL_46 | ···················· | ···················· | ···················· | ···················· | ······NNNNNNNN······ | ···················· | 1 |
| D | 5183 | hCoV-19/Switzerland/BE-ETHZ-100059/2020\|EPI_ISL_46 | ···················· | ···················· | ···················· | ···················· | ······NNNNNNNNNNNNNN | ···················· | 1 |
| D | 5184 | hCoV-19/Switzerland/VD-ETHZ-100060/2020\|EPI_ISL_46 | ···················· | ···················· | ···················· | ···················· | ······NNNNNNNNNNNNNN | NNNNNNNNNNNNNNNNNNNN | 1 |
| D | 5185 | hCoV-19/Switzerland/GR-ETHZ-100061/2020\|EPI_ISL_46 | ···················· | ···················· | ···················· | ···················· | ······NNNNNNNNNNNNNN | NNNNNNNNNNNNNNNNNNNN | 1 |
| D | 5186 | hCoV-19/Switzerland/SZ-ETHZ-100062/2020\|EPI_ISL_46 | ···················· | ···················· | ···················· | ···················· | ······NNNNNNNNNNNNNN | NNNNNNNNNNNNNNNNNNNN | 1 |
| D | 5187 | hCoV-19/Switzerland/BS-ETHZ-100063/2020\|EPI_ISL_46 | ···················· | ···················· | ···················· | ···················· | ······NNNNNNNNNNNNNN | NNNNNNNNNNNNNNNNNNNN | 1 |
| D | 5188 | hCoV-19/Switzerland/SZ-ETHZ-100064/2020\|EPI_ISL_46 | ···················· | ···················· | ···················· | ···················· | ······NNNNNNNNNNNNNN | ···················· | 1 |
| D | 5189 | hCoV-19/Switzerland/GR-ETHZ-100065/2020\|EPI_ISL_46 | ···················· | ···················· | ···················· | ···················· | ······NNNNNNNNNNNNNN | NNNNNNNNNNNNNNNNNNNN | 1 |
| D | 5190 | hCoV-19/Switzerland/GR-ETHZ-100066/2020\|EPI_ISL_46 | ···················· | ···················· | ···················· | ···················· | ······NNNNNNNNNNNNNN | W··················· | 1 |
| D | 5191 | hCoV-19/Switzerland/ZG-ETHZ-100067/2020\|EPI_ISL_46 | ···················· | ···················· | ···················· | ···················· | ······NNNNNNNNNNNNNN | NNNNNNNNNNNNNNNNNNNN | 1 |
| D | 5192 | hCoV-19/Switzerland/TI-ETHZ-100068/2020\|EPI_ISL_46 | ···················· | ···················· | ···················· | ···················· | ······NNNNNNNNNNNNNN | NNNNNNNNNNNNNNNNNNNN | 1 |

```
D 5194 hCoV-19/Switzerland/TI-ETHZ-100070/2020|EPI_ISL_46 ···················· ···················· ···················· ···················· ·······NNNNNNNNNNNNN ···················· 1
D 5211 hCoV-19/Singapore/376/2020|EPI_ISL_469125|2020-04- ···················· ···················· ···················· ···················· NNNNNNNNNNNNNNNNNNNN NNNNNNNNNNNNNNNNNNNN 1
D 5214 hCoV-19/Singapore/379/2020|EPI_ISL_469128|2020-04- ···················· ···················· ···················· ···················· NNNN················ ···················· 1
D 5411 hCoV-19/England/201061439/2020|EPI_ISL_464410|2020 ···················· ···················· ···················· ···················· ···················· NNNNNNNNNNNN·NN····· 1
D 5413 hCoV-19/England/201080072/2020|EPI_ISL_464457|2020 ···················· ···················· ···················· ···················· ··N···NNNNNNNNNNNNN· NNNNN··············· 1
D 5414 hCoV-19/England/20108007504/2020|EPI_ISL_464458|20 ···················· ···················· ···················· ···················· ·NNNNNNNNNNNNNNNNNNN NNNNNNNNNNNNNNNNNNNN 1
D 5420 hCoV-19/England/201080082/2020|EPI_ISL_464465|2020 ···················· ···················· ···················· ···················· NN·NNNNNNN··NNNNNN·N N·NNN····NN··NNNN·NN 1
D 5423 hCoV-19/England/201080092/2020|EPI_ISL_464468|2020 ···················· ···················· ···················· ···················· ·NNNNNNN········NN·N NNNNNNNNNNNNNNNNN··· 1
D 5424 hCoV-19/England/201080162/2020|EPI_ISL_464472|2020 ···················· ···················· ···················· ···················· NNNNNNNNNNNNNNNNNNNN N·NNNNNNNNNNN·N·N··· 1
D 5427 hCoV-19/England/201081131/2020|EPI_ISL_464476|2020 ···················· ···················· ···················· ···················· ·NNNNNNNNNNNNNNNNNNN ···················· 1
D 5433 hCoV-19/England/20108152504/2020|EPI_ISL_464485|20 ···················· ···················· ···················· ···················· ·NNNNNNNNNNNNNNNN··· NNNNNNNNNNNNNNNNNNNN 1
D 5439 hCoV-19/England/20122017304/2020|EPI_ISL_465739|20 ···················· ···················· ···················· ···················· ···················· N···N····NNNNNNNNNNN 1
D 5445 hCoV-19/Singapore/346/2020|EPI_ISL_469101|2020-04- ···················· ···················· ···················· ···················· ···················· ··········NNNNN····· 1
D 5474 hCoV-19/DRC/4026/2020|EPI_ISL_471404|2020-04-27 ···················· ···················· ···················· ···················· NNNNNNNNNNNNNNNNNNNN NNNNNNNNNNNNNNN····· 1
D 5521 hCoV-19/Wales/PHWC-15E0B7/2020|EPI_ISL_472792|2020 ···················· ···················· ···················· ···················· NNNNNNNNNNNNNNNNNNNN NNNNNNNNNNNNNNNNNNNN 1
D 5600 hCoV-19/USA/NE-10832/2020|EPI_ISL_475184|2020-05-0 ···················· ···················· ···················· ···················· NNNNNNNNNNNNNNNNNNNN NNNNNNNNNNNNNNNNNNNN 1
D 5656 hCoV-19/England/201060300/2020|EPI_ISL_464401|2020 ···················· ···················· ···················· ···················· ·NNNNNNNNNNN········ ···················· 1
D 5657 hCoV-19/England/201061225/2020|EPI_ISL_464408|2020 ···················· ···················· ···················· ···················· ·NNNNNNNNNNNNNNNNNNN ···················· 1
D 5660 hCoV-19/England/201061455/2020|EPI_ISL_464413|2020 ···················· ···················· ···················· ···················· ···················· N·NNN····N·········· 1
D 5662 hCoV-19/England/201061458/2020|EPI_ISL_464416|2020 ···················· ···················· ···················· ···················· NNNNNNNNNNNNNNNNNNNN NNNNNNNNNNNNNNNNNNNN 1
D 5664 hCoV-19/England/201080004/2020|EPI_ISL_464419|2020 ···················· ···················· ···················· ···················· ·····N·············· NNNNNNNNNNNNNNNNNNNN 1
D 5665 hCoV-19/England/201080005/2020|EPI_ISL_464420|2020 ···················· ···················· ···················· ···················· NNNNNNNNNNNNNNNNNNNN NNNNNNNNNNNNNNNNNNNN 1
D 5670 hCoV-19/England/201080014/2020|EPI_ISL_464426|2020 ···················· ···················· ···················· ···················· NNNNNNNNNNNNNNNNNNNN NNNNNNNNNNNNNNNNNNNN 1
D 5672 hCoV-19/England/201080021/2020|EPI_ISL_464428|2020 ···················· ···················· ···················· ···················· ·NNNNNNNNNNNNNNNNNNN NNNNNNNNNNNNNNNNNNNN 1
D 5673 hCoV-19/England/201080022/2020|EPI_ISL_464429|2020 ···················· ···················· ···················· ···················· NNNNNNNNNNNNNNNNNNNN NNNNNNNNNNNNNNNNNNNN 1
D 5674 hCoV-19/England/201080024/2020|EPI_ISL_464430|2020 ···················· ···················· ···················· ···················· NNNNNNNNNNNNNNNNNNNN NNNNNNNNNNNNNNNNNNNN 1
D 5677 hCoV-19/England/201080028/2020|EPI_ISL_464433|2020 ···················· ···················· ···················· ···················· ·NNNNNNNNNNNNNNNNNNN NNNNNNNNNNNNNNNNNNNN 1
D 5679 hCoV-19/England/201080031/2020|EPI_ISL_464435|2020 ···················· ···················· ···················· ···················· NNNNNNNNNNNNNNNNNNNN NNNNNNNNNNNNNNNNNNNN 1
D 5680 hCoV-19/England/201080038/2020|EPI_ISL_464437|2020 ···················· ···················· ···················· ···················· ···················· NNNNNNNNNNNNNNNNNNNN 1
D 5682 hCoV-19/England/201080042/2020|EPI_ISL_464439|2020 ···················· ···················· ···················· ···················· NNNNNNNNNNNNNNNNNNNN NNNNNNNNNNNNNNN·NN·· 1
D 5683 hCoV-19/England/201080043/2020|EPI_ISL_464440|2020 ···················· ···················· ···················· ···················· NNNNNNNNNNNNNNNNNNNN ····N··············· 1
D 5688 hCoV-19/England/201080055/2020|EPI_ISL_464446|2020 ···················· ···················· ···················· ···················· ·N··NN·NNN·NNN··NNNN NNNNNNNNNNNNNNNNNNNN 1
D 5690 hCoV-19/England/201080057/2020|EPI_ISL_464448|2020 ···················· ···················· ···················· ···················· NNNNNNNNNNNNNNNNNNNN NNNNNNNNNNNNNNN·N··· 1
D 5693 hCoV-19/England/201080063/2020|EPI_ISL_464451|2020 ···················· ···················· ···················· ···················· NNNNNNNN·NNNN·N··N·N NNNNNNNNNNNN·NN····· 1
D 5709 hCoV-19/England/20120054704/2020|EPI_ISL_465725|20 ···················· ···················· ···················· ···················· NNNNNNNNNNNNNNNNNNNN NNNNNNNNNNNNNNNNNNNN 1
D 5713 hCoV-19/England/20122017904/2020|EPI_ISL_465742|20 ···················· ···················· ···················· ···················· NNNNNNNNNNNNNNNNNNNN ···················N 1
D 5714 hCoV-19/England/20122030404/2020|EPI_ISL_465748|20 ···················· ···················· ···················· ···················· ·····N····NNNN··N··· N··NN·N··N·········· 1
D 5715 hCoV-19/England/20122031904/2020|EPI_ISL_465753|20 ···················· ···················· ···················· ···················· ··NN·N·NN··········· NNN·N·N·N·······N··· 1
D 5718 hCoV-19/England/20122069804/2020|EPI_ISL_465759|20 ···················· ···················· ···················· ···················· ···················· ·NNNNN······N······· 1
D 5721 hCoV-19/England/20122121504/2020|EPI_ISL_465768|20 ···················· ···················· ···················· ···················· ·NNNNNNNNNNNNNNNNNNN NNNNNNNNNNNNNNNNNNNN 1
D 5724 hCoV-19/England/20124010904/2020|EPI_ISL_465780|20 ···················· ···················· ···················· ···················· ···················· NNNNNNNNNNNNNNN·N··· 1
D 6009 hCoV-19/England/20238073704/2020|EPI_ISL_471515|20 ···················· ···················· ···················· ···················· ··NNNNNNNNNNNNNNNNNN NNNNNNNNNNNNNNNNNNNN 1
D 6012 hCoV-19/England/20240010804/2020|EPI_ISL_471518|20 ···················· ···················· ···················· ···················· ···················· NNNNNNNNNNNNNNNNNNNN 1
D 6013 hCoV-19/England/20242082604/2020|EPI_ISL_471520|20 ···················· ···················· ···················· ···················· ·NNNNNNNN··N·NNNN··· NNNN··········N····· 1
D 6016 hCoV-19/England/20246148004/2020|EPI_ISL_471527|20 ···················· ···················· ···················· ···················· ·NNNNNNNNNNNNNNNNNNN NNNNNNNNNNNNNNNNNNNN 1
D 6065 hCoV-19/Belgium/UGent-111/2020|EPI_ISL_475071|2020 ···················· ···················· ···················· ···················· NNNNNNNNNNNNNNNNNNNN ···················· 1
D 6069 hCoV-19/Belgium/UGent-115/2020|EPI_ISL_475075|2020 ···················· ···················· ···················· ···················· NNNNNNNNNNNNNNNNNNNN NNNNNNNNNNNNNNNNNNNN 1
D 6072 hCoV-19/Belgium/UGent-118/2020|EPI_ISL_475078|2020 ···················· ···················· ···················· ···················· NNNNNNNNNNNNNNNNNNNN NNNNNNNNNNNNNNNNNNNN 1
D 6073 hCoV-19/Belgium/UGent-119/2020|EPI_ISL_475079|2020 ···················· ···················· ···················· ···················· NNNNNNNNNNNNNNNNNNNN NNNNNNNNNNNNNNNNNNNN 1
D 6074 hCoV-19/Belgium/UGent-120/2020|EPI_ISL_475080|2020 ···················· ···················· ···················· ···················· NNNNNNNNNNNNNNNNNNNN NNNNNNNNNNNNNNNNNNNN 1
D 6075 hCoV-19/Belgium/UGent-121/2020|EPI_ISL_475081|2020 ···················· ···················· ···················· ···················· NNNNNNNNNNNNNNNNNNNN NNNNNNNNNNNNNNNNNNNN 1
D 6094 hCoV-19/Switzerland/VD-ETHZ-140067/2020|EPI_ISL_47 ···················· ···················· ···················· ···················· ···················· ······MMKKK·M·MKWMKM 1
D 6107 hCoV-19/Switzerland/AG-ETHZ-140086/2020|EPI_ISL_47 ···················· ···················· ···················· ···················· ·······NNNNNNNNNNNNN ···················· 1
D 6170 hCoV-19/England/20180011204/2020|EPI_ISL_465428|20 ···················· ···················· ···················· ···················· ·NNNNNNNNNNNNNNNNNNN NNNNNNNNNNNNNNNNNNNN 1
D 6174 hCoV-19/England/20186075804/2020|EPI_ISL_465474|20 ···················· ···················· ···················· ···················· NNNNNNNNNNNNNNNNNNNN NNNNNNNNNNNNNNNNNNNN 1
D 6178 hCoV-19/England/20186097004/2020|EPI_ISL_465482|20 ···················· ···················· ···················· ···················· ·NNNNNNNNNNNNNNNNNNN NNNNNNNNNNNNNNNNNNNN 1
D 6179 hCoV-19/England/20186106204/2020|EPI_ISL_465485|20 ···················· ···················· ···················· ···················· NNNNNNNNNNNNNNNNNNNN NNNNNNNNNNNNNNNNNNNN 1
D 6182 hCoV-19/England/20188012604/2020|EPI_ISL_465492|20 ···················· ···················· ···················· ···················· NNNNNNNNNNNNNNNNNNNN NN···NN·····N······· 1
D 6320 hCoV-19/Switzerland/VD-ETHZ-140052/2020|EPI_ISL_47 ···················· ·A·················· ···················· ···················· ·······SNNNNNNNNNNNN ···················· 1
D 6327 hCoV-19/Switzerland/LU-ETHZ-140064/2020|EPI_ISL_47 ···················· ···················· ···················· ···················· ·······NNNNNNNNNNNNN ···················· 1
D 6355 hCoV-19/England/NORW-ED1AC/2020|EPI_ISL_478720|202 ···················· ···················· ···················· ···················· NNNNNNNNNNNNNNNNNNNN NNNNNNNNNNNNNNNNNNNN 1
D 6437 hCoV-19/England/20099069904/2020|EPI_ISL_464216|20 ···················· ···················· ···················· ···················· ···N·N··NNNN······NN ···················· 1
D 6441 hCoV-19/England/20099072104/2020|EPI_ISL_464222|20 ···················· ···················· ···················· ···················· ·····N······N······· NNNNNNNNNNNNNNNNNN·N 1
D 6456 hCoV-19/England/20189155001/2020|EPI_ISL_465551|20 ···················· ···················· ···················· ···················· ·········N···NN····N NNNNNNNNNNNNNNNNNNNN 1
D 6467 hCoV-19/England/20189159601/2020|EPI_ISL_465564|20 ···················· ···················· ···················· ···················· NNNNNNNNNNNNNNNNNNNN ··········N········· 1
D 6470 hCoV-19/England/20189166001/2020|EPI_ISL_465569|20 ···················· ···················· ···················· ···················· ···················· NNNN················ 1
D 6475 hCoV-19/England/20189183004/2020|EPI_ISL_465579|20 ···················· ···················· ···················· ···················· ·NNNNNNNNNNNNNNNNNNN ··NN················ 1
D 6480 hCoV-19/England/20190038701/2020|EPI_ISL_465584|20 ···················· ···················· ···················· ···················· ···················· NNNNNNNNNNNNNNNN···· 1
D 6499 hCoV-19/Germany/BY-MVP-0240/2020|EPI_ISL_466899|20 ···················· ···················· ···················· ···················· NNNNNNNNNNNNNNNNNNNN ···················· 1
D 6580 hCoV-19/Scotland/EDB5818/2020|EPI_ISL_473861|2020- ···················· ···················A ···················· ···················· NNNNNNNNNNNNNNNNNNNN NNNNNNNNNNNNNNNNNNNN 1
D 6585 hCoV-19/Scotland/EDB5882/2020|EPI_ISL_473866|2020- ···················· ···················A ···················· ···················· NNNNNNNNNNNNNNNNNNNN NNNNNNNNNNNNNNNNNNNN 1
D 6591 hCoV-19/Scotland/EDB6048/2020|EPI_ISL_473872|2020- ···················· ···················A ···················· ···················· NNNNNNNNNNNNNNNNNNNN NNNNNNNNNNNNNNNNNNNN 1
D 6651 hCoV-19/Switzerland/ZH-ETHZ-140042/2020|EPI_ISL_47 ···················· ···················· ···················· ···················· ·······NNNNNNNNNNNNN ···················· 1
D 6654 hCoV-19/Switzerland/BE-ETHZ-140048/2020|EPI_ISL_47 ···················· ·A·················· ···················· ···················· ·······NNNNNNNNNNNNN ···················· 1
D 6662 hCoV-19/England/20099048604/2020|EPI_ISL_464207|20 ···················· ···················· ···················· ···················· ·NNNNNNNNNNNNN······ ···················· 1
D 6663 hCoV-19/England/20099066104/2020|EPI_ISL_464208|20 ···················· ···················· ···················· ···················· ···················· NNNNN·NN············ 1
D 6667 hCoV-19/England/20100078004/2020|EPI_ISL_464258|20 ···················· ···················· ···················· ···················· NNNNNNNNNNNNNNNNNNNN NNNNNNNNNNNNNNNNNNNN 1
D 6668 hCoV-19/England/20102000402/2020|EPI_ISL_464260|20 ···················· ···················· ···················· ···················· NNNNNNNNNNNNNNNNNNNN NNNNNNNNNNNNNNNNNNNN 1
D 6670 hCoV-19/England/20102001304/2020|EPI_ISL_464266|20 ···················· ···················· ···················· ···················· NNNNNNNNNNNNNNNNNNNN ··NNNNNNNNNNNNNNNNNN 1
D 6675 hCoV-19/England/20102014904/2020|EPI_ISL_464288|20 ···················· ···················· ···················· ···················· ··NN···NNNNN·······N ···················· 1
```

| | | | | | | | | | |
|---|---|---|---|---|---|---|---|---|---|
| D | 6677 | hCoV-19/England/201021553/2020\|EPI_ISL_464292\|2020 | ···················· | ···················· | ···················· | ···················· | NNNNNNNNNNNNNNNNNNNN | NNNNNNNNNNNNNNNNNNNN | 1 |
| D | 6683 | hCoV-19/England/201021561/2020\|EPI_ISL_464298\|2020 | ···················· | ···················· | ···················· | ···················· | NNNNNNNNNNNNNNNNNNNN | NNNNNNNNNNNNNNNNNNNN | 1 |
| D | 6684 | hCoV-19/England/20188027704/2020\|EPI_ISL_465500\|20 | ···················· | ···················· | ···················· | ···················· | NNNNNNNNNNNNNNNNNNNN | NNNNNNNNNNNNNNNNNNNN | 1 |
| D | 6685 | hCoV-19/England/20188033004/2020\|EPI_ISL_465501\|20 | ···················· | ···················· | ···················· | ···················· | ··NNNNNNNNNNNNNNNNNN | NNNN·············NNN | 1 |
| D | 6686 | hCoV-19/England/20188034704/2020\|EPI_ISL_465503\|20 | ···················· | ···················· | ···················· | ···················· | NNNNNNNNNNNNNNNNNNNN | NNNNNNNNNNNNNNNNNNNN | 1 |
| D | 6688 | hCoV-19/England/20188036404/2020\|EPI_ISL_465507\|20 | ···················· | ···················· | ···················· | ···················· | NNNNNNNNNNNNNNNNNNNN | NNNNNNNNNNNNNNNNNNNN | 1 |
| D | 6694 | hCoV-19/England/20188145804/2020\|EPI_ISL_465519\|20 | ···················· | ···················· | ···················· | ···················· | ···NNNNN·····NN·NN·N | NN·················· | 1 |
| D | 6695 | hCoV-19/England/20188152504/2020\|EPI_ISL_465520\|20 | ···················· | ···················· | ···················· | ···················· | NNNNNNNNNNNNNNNNNNNN | NNNNNNNNNNNNNNNNNNNN | 1 |
| D | 6696 | hCoV-19/England/20188153704/2020\|EPI_ISL_465521\|20 | ···················· | ···················· | ···················· | ···················· | NNNNNNNNNNNNNNNNNNNN | NNNNNNNNNNNNNNNNNNNN | 1 |
| D | 6697 | hCoV-19/England/20188160304/2020\|EPI_ISL_465523\|20 | ···················· | ···················· | ···················· | ···················· | ·NNNNNNNNNNNNNNNNNNN | ···················· | 1 |
| D | 6700 | hCoV-19/England/20189042401/2020\|EPI_ISL_465527\|20 | ···················· | ···················· | ···················· | ···················· | NNNNNNNNNNNNNNNNNNNN | NNNNNNNNNNNNNNNNNNNN | 1 |
| D | 6701 | hCoV-19/England/20189042501/2020\|EPI_ISL_465528\|20 | ···················· | ···················· | ···················· | ···················· | ···················· | NNNNNNNNNNNNNNNNNNNN | 1 |
| D | 6703 | hCoV-19/England/20189049401/2020\|EPI_ISL_465532\|20 | ···················· | ···················· | ···················· | ···················· | NNNN················ | NNNN·········N·····N | 1 |
| D | 6706 | hCoV-19/England/20189054201/2020\|EPI_ISL_465536\|20 | ···················· | ···················· | ···················· | ···················· | NNNNNNNNNNNNNNNNNNNN | NNNNNNNNNNNNNNNNNNNN | 1 |
| D | 6709 | hCoV-19/England/20189118901/2020\|EPI_ISL_465543\|20 | ···················· | ···················· | ···················· | ···················· | ··NNNNNNNNNNNNNNNNNN | NNNN················ | 1 |
| D | 6711 | hCoV-19/England/20189148201/2020\|EPI_ISL_465549\|20 | ···················· | ···················· | ···················· | ···················· | NNNNNNNNNNNNNNNNNNNN | NNNNNNNNNNNNNNNNNNNN | 1 |
| D | 6714 | hCoV-19/England/20190072401/2020\|EPI_ISL_465591\|20 | ···················· | ···················· | ···················· | ···················· | NNNNNNNNNNNNNNNNNNNN | NNNNNNNNNNNNNNNNNNNN | 1 |
| D | 6716 | hCoV-19/England/20190142001/2020\|EPI_ISL_465593\|20 | ···················· | ···················· | ···················· | ···················· | NNNNNNNNNNNNNNNNNNNN | NNNNNNNNNNNNNNNNNNNN | 1 |
| D | 6717 | hCoV-19/England/20192003004/2020\|EPI_ISL_465594\|20 | ···················· | ···················· | ···················· | ···················· | NNNNNNNNNNNNNNNNNNNN | N·N·NN·············· | 1 |
| D | 6719 | hCoV-19/England/20192009801/2020\|EPI_ISL_465596\|20 | ···················· | ···················· | ···················· | ···················· | ·NNNNNNNNNNNNNNNNNNN | NNNNNNNNNNNNNNNNNNNN | 1 |
| D | 6943 | hCoV-19/England/20096051104/2020\|EPI_ISL_464188\|20 | ···················· | ···················· | ···················· | ···················· | ·NNNNNNNNNNNNNNNNNNN | NNNNN···NN·········· | 1 |
| D | 6954 | hCoV-19/Spain/IB-IBV-2522/2020\|EPI_ISL_469000\|2020 | ···················· | ···················· | ···················· | ···················· | NNNNNNNNNNNNNNNNNNNN | NNNNNNNNNNNNNNNNNNNN | 1 |
| D | 6955 | hCoV-19/Spain/IB-IBV-2512/2020\|EPI_ISL_469001\|2020 | ···················· | ···················· | ···················· | ···················· | NNNNNNNNNNNNNNNNNNNN | NNNNNNNNNNNNNNNNNNNN | 1 |
| D | 6956 | hCoV-19/Spain/IB-IBV-2532/2020\|EPI_ISL_469003\|2020 | ···················· | ···················· | ···················· | ···················· | NNNNNNNNNNNNNNNNNNNN | NNNNN···NNNNNNNNNNNN | 1 |
| D | 6957 | hCoV-19/Spain/IB-IBV-2507/2020\|EPI_ISL_469004\|2020 | ···················· | ···················· | ···················· | ···················· | NNNNNNNNNNNNNNNNNNNN | NNNNNNNNNNNNNNNNNNNN | 1 |
| D | 6958 | hCoV-19/Spain/IB-IBV-2570/2020\|EPI_ISL_469006\|2020 | ···················· | ···················· | ···················· | ···················· | NNNNNNNNNNNNNNNNNNNN | NNNNNNNNNNNNNNNNNNNN | 1 |
| D | 6960 | hCoV-19/Spain/IB-IBV-2552/2020\|EPI_ISL_469009\|2020 | ···················· | ···················A | ···················· | ···················· | NNNNNNNNNNNNNNNNNNNN | NNNNNNNNNNNNNNNNNNNN | 1 |
| D | 6961 | hCoV-19/Spain/IB-IBV-2516/2020\|EPI_ISL_469010\|2020 | ···················· | ···················· | ···················· | ···················· | NNNNNNNNNNNNNNNNNNNN | ···················· | 1 |
| D | 6962 | hCoV-19/Spain/IB-IBV-2526/2020\|EPI_ISL_469012\|2020 | ···················· | ···················· | ···················· | ···················· | NNNNNNNNNNNNNNNNNNNN | NNNNNNNNNNNNNNNNNNNN | 1 |
| D | 6963 | hCoV-19/Spain/IB-IBV-2511/2020\|EPI_ISL_469013\|2020 | ···················· | ···················· | ···················· | ···················· | NNNNNNNNNNNNNNNNNNNN | NNNNNNNNNNNNNNNNNNNN | 1 |
| D | 6964 | hCoV-19/Spain/IB-IBV-2525/2020\|EPI_ISL_469014\|2020 | ···················· | ···················· | ···················· | ···················· | NNNNNNNNNNNNNNNNNNNN | NNNNNNNNNNNNNNNNNNNN | 1 |
| D | 7136 | hCoV-19/Wales/PHWC-163556/2020\|EPI_ISL_473961\|2020 | ···················· | ···················· | ···················· | ···················· | NNNN················ | ···················· | 1 |
| D | 7262 | hCoV-19/England/201140405/2020\|EPI_ISL_464732\|2020 | ···················· | ···················· | ···················· | ···················· | ·······NNNNNNNNNNNNN | NNNNNNNNNNNNNNNNNNNN | 1 |
| D | 7264 | hCoV-19/England/201140442/2020\|EPI_ISL_464735\|2020 | ···················· | ···················· | ···················· | ···················· | NNNNNNNNNNNNNNNNNNNN | NNNNNNNNNNNNNNNNNNNN | 1 |
| D | 7265 | hCoV-19/England/201140586/2020\|EPI_ISL_464738\|2020 | ···················· | ···················· | ···················· | ···················· | ·NNNNNNNNNN·N······· | NNNNNNNNNNNNNNNNNNNN | 1 |
| D | 7266 | hCoV-19/England/201140778/2020\|EPI_ISL_464741\|2020 | ···················· | ···················· | ···················· | ···················· | ·N·················· | NNNN················ | 1 |
| D | 7269 | hCoV-19/England/201141169/2020\|EPI_ISL_464744\|2020 | ···················· | ···················· | ···················· | ···················· | ··NNN·········NN···· | ··············NN···· | 1 |
| D | 7271 | hCoV-19/England/201141357/2020\|EPI_ISL_464751\|2020 | ···················· | ···················· | ···················· | ···················· | ·NNNNNNNNNNNNNNNNNNN | NNNNNNNNNNNNNNNNNNNN | 1 |
| D | 7273 | hCoV-19/England/201141519/2020\|EPI_ISL_464757\|2020 | ···················· | ···················· | ···················· | ···················· | ·NNNNNNNNNNNNNNNNNNN | NNNNNNNNNNNNNNNNNNNN | 1 |
| D | 7275 | hCoV-19/England/201141521/2020\|EPI_ISL_464759\|2020 | ···················· | ···················· | ···················· | ···················· | ·NNNNNNNNNNNNNNNNNNN | NNNNNNNNNNNNNNNNNNNN | 1 |
| D | 7276 | hCoV-19/England/201141523/2020\|EPI_ISL_464761\|2020 | ···················· | ···················· | ···················· | ···················· | NNNNNNNNNNNNNNNNNNNN | NNNNNNNNNNNNNNNNNNNN | 1 |
| D | 7279 | hCoV-19/England/201141660/2020\|EPI_ISL_464769\|2020 | ···················· | ···················· | ···················· | ···················· | ·NNNNNNNNNNNNNNNNNNN | NNNNNNNNNNNNNNNNNNNN | 1 |
| D | 7281 | hCoV-19/England/201160003/2020\|EPI_ISL_464771\|2020 | ···················· | ···················· | ···················· | ···················· | NNNNNNNNNNNNNNNNNNNN | NNNNNNNNNNNNNNNNNNNN | 1 |
| D | 7282 | hCoV-19/England/201160004/2020\|EPI_ISL_464772\|2020 | ···················· | ···················· | ···················· | ···················· | NNNNNNNNNNNNNNNNNNNN | NNNNNNNNNNNNNNNNNNNN | 1 |
| D | 7331 | hCoV-19/Wales/PHWC-164160/2020\|EPI_ISL_474135\|2020 | ···················· | ···················· | ···················· | ···················· | ·····NNNNNNNNNNNNNNN | NNNNNNNNNNNNNNNNNNNN | 1 |
| D | 7450 | hCoV-19/England/201140250/2020\|EPI_ISL_464715\|2020 | ···················· | ···················· | ···················· | ···················· | ···················· | NNNNNNNNNNNNNNNNN··· | 1 |
| D | 7451 | hCoV-19/England/201140293/2020\|EPI_ISL_464720\|2020 | ···················· | ···················· | ···················· | ···················· | ··NNNNNNNNNN···NNNN· | ··········N······NNN | 1 |
| D | 7455 | hCoV-19/England/201160007/2020\|EPI_ISL_464774\|2020 | ···················· | ···················· | ···················· | ···················· | NNNNNNNNNNNNNNNNNNNN | NNNNNNNNNNNNNNNNNNNN | 1 |
| D | 7459 | hCoV-19/England/201160037/2020\|EPI_ISL_464779\|2020 | ···················· | ···················· | ···················· | ···················· | ··············N····· | NNNNNNNNNNNNNNNNNNNN | 1 |
| D | 7460 | hCoV-19/England/201160048/2020\|EPI_ISL_464780\|2020 | ···················· | ···················· | ···················· | ···················· | NNNNNNNNNNNNNNNNNNNN | NNNNNNNNNNNNNNNNNNNN | 1 |
| D | 7466 | hCoV-19/England/201160093/2020\|EPI_ISL_464786\|2020 | ···················· | ···················· | ···················· | ···················· | ···················· | NNNNNNNNNNNNNNNNN··· | 1 |
| D | 7468 | hCoV-19/England/201160217/2020\|EPI_ISL_464789\|2020 | ···················· | ···················· | ···················· | ···················· | ···················· | NNNNNNNNN··NNNN····· | 1 |
| D | 7471 | hCoV-19/England/201160261/2020\|EPI_ISL_464792\|2020 | ···················· | ···················· | ···················· | ···················· | NNNNNNNNNNNNN···N··· | NNNN················ | 1 |
| D | 7473 | hCoV-19/England/201160274/2020\|EPI_ISL_464794\|2020 | ···················· | ···················· | ···················· | ···················· | ·NNNNNNNNNNNNNNNNNNN | ···················· | 1 |
| D | 7475 | hCoV-19/England/201160623/2020\|EPI_ISL_464796\|2020 | ···················· | ···················· | ···················· | ···················· | NNNNNNNNNNNNNNNNNNNN | NNNNNNNNNNNNNNNNNNNN | 1 |
| D | 7477 | hCoV-19/England/201160629/2020\|EPI_ISL_464798\|2020 | ···················· | ···················· | ···················· | ···················· | ·NNNNNNNNNNNNNNNNNNN | NNNNNN·NNN·NNNNN···· | 1 |
| D | 7478 | hCoV-19/England/201160641/2020\|EPI_ISL_464799\|2020 | ···················· | ···················· | ···················· | ···················· | ·NNNNNNNNNNNNNNNNNNN | ·········R·········· | 1 |
| D | 7480 | hCoV-19/Switzerland/ZH-ETHZ-120006/2020\|EPI_ISL_46 | ···················· | ·A·················· | ···················· | ···················· | ·······NNNNNNNNNNNNN | NNNNNNNNNNNNNNNNNNNN | 1 |
| D | 7481 | hCoV-19/Switzerland/GE-ETHZ-120007/2020\|EPI_ISL_46 | ···················· | ···················· | ···················· | ···················· | ·······NNNNNNNNNNNNN | ···················· | 1 |
| D | 7482 | hCoV-19/Switzerland/BS-ETHZ-120014/2020\|EPI_ISL_46 | ···················· | ···················· | ···················· | ···················· | ·······NNNNNNNNNNNNN | NNNNNNNNNNNNNNNNNNNN | 1 |
| D | 7483 | hCoV-19/Switzerland/BL-ETHZ-120044/2020\|EPI_ISL_46 | ···················· | ···················· | ···················· | ···················· | ·······NNNNNNNNNNNNN | ···················· | 1 |
| D | 7484 | hCoV-19/Switzerland/ZH-ETHZ-120045/2020\|EPI_ISL_46 | ···················· | ···················· | ···················· | ···················· | ·······NNNNNNNNNNNNN | ···················· | 1 |
| D | 7485 | hCoV-19/Switzerland/BE-ETHZ-120068/2020\|EPI_ISL_46 | ···················· | ···················· | ···················· | ···················· | ·······NNNNNNNNNNNNN | NNNNNNNNNNNNNNNNNNNN | 1 |
| D | 7486 | hCoV-19/Switzerland/BE-ETHZ-120070/2020\|EPI_ISL_46 | ···················· | ···················· | ···················· | ···················· | ·······NNNNNNNNNNNNN | NNNNNNNNNNNNNNNNNNNN | 1 |
| D | 7488 | hCoV-19/Switzerland/GR-ETHZ-120166/2020\|EPI_ISL_46 | ···················· | ···················· | ···················· | ···················· | ·······NNNNNNNNNNNNN | NNNNNNNNNNNNNNNNNNNN | 1 |
| D | 7490 | hCoV-19/Switzerland/UR-ETHZ-120170/2020\|EPI_ISL_46 | ···················· | ···················· | ···················· | ···················· | ·······NNNNNNNNNNNNN | NNNNNNNNNNNNNNNNNNNN | 1 |
| D | 7491 | hCoV-19/Switzerland/AG-ETHZ-120172/2020\|EPI_ISL_46 | ···················· | ···················· | ···················· | ···················· | ·······NNNNNNNNNNNNN | ···················· | 1 |
| D | 7493 | hCoV-19/Switzerland/VD-ETHZ-120179/2020\|EPI_ISL_46 | ···················· | ···················· | ···················· | ···················· | ·······NNNNNNNNNNNNN | NNNNNNNNNNNNNNNNNNNN | 1 |
| D | 7495 | hCoV-19/Switzerland/UR-ETHZ-120188/2020\|EPI_ISL_46 | ···················· | ···················· | ···················· | ···················· | ·······NNNNNNNNNNNNN | NNNNNNNNNNNNNNNNNNNN | 1 |
| D | 7496 | hCoV-19/Switzerland/ZH-ETHZ-120191/2020\|EPI_ISL_46 | ···················· | ···················· | ···················· | ···················· | ·······NNNNNNNNNNNNN | NNNNNNNNNNNNNNNNNNNN | 1 |
| D | 7497 | hCoV-19/Switzerland/ZH-ETHZ-120194/2020\|EPI_ISL_46 | ···················· | ···················· | ···················· | ···················· | ·······NNNNNNNNNNNNN | NNNNNNNNNNNNNNNNNNNN | 1 |
| D | 7499 | hCoV-19/Switzerland/TI-ETHZ-120203/2020\|EPI_ISL_46 | ···················· | ···················· | ···················· | ···················· | ·······NNNNNNNNNNNNN | NNNNNNNNNNNNNNNNNNNN | 1 |
| D | 7500 | hCoV-19/Switzerland/ZH-ETHZ-120206/2020\|EPI_ISL_46 | ···················· | ···················· | ···················· | ···················· | ·······NNNNNNNNNNNNN | NNNNNNNNNNNNNNNNNNNN | 1 |
| D | 7501 | hCoV-19/Switzerland/BL-ETHZ-120207/2020\|EPI_ISL_46 | ···················· | ···················· | ···················· | ···················· | ·······NNNNNNNNNNNNN | NNNNNNNNNNNNNNNNNNNN | 1 |
| D | 7502 | hCoV-19/Switzerland/GE-ETHZ-120219/2020\|EPI_ISL_46 | ···················· | ·R·················· | ···················· | ···················· | ·······NNNNNNNNNNNNN | NNNNNNNNNNNNNNNNNNNN | 1 |
| D | 7504 | hCoV-19/Switzerland/BE-ETHZ-120223/2020\|EPI_ISL_46 | ···················· | ·R·················· | ···················· | ···················· | ·······NNNNNNNNNNNNN | NNNNNNNNNNNNNNNNNNNN | 1 |
| D | 7506 | hCoV-19/Switzerland/UR-ETHZ-120232/2020\|EPI_ISL_46 | ···················· | ···················· | ···················· | ···················· | ·······NNNNNNNNNNNNN | NNNNNNNNNNNNNNNNNNNN | 1 |
| D | 7507 | hCoV-19/Switzerland/BE-ETHZ-120237/2020\|EPI_ISL_46 | ···················· | ···················· | ···················· | ···················· | ·······NNNNNNNNNNNNN | ····W··············· | 1 |
| D | 7508 | hCoV-19/Switzerland/BE-ETHZ-120238/2020\|EPI_ISL_46 | ···················· | ···················· | ···················· | ···················· | ·······NNNNNNNNNNNNN | NNNNNNNNNNNNNNNNNNNN | 1 |
| D | 7746 | hCoV-19/England/201161700/2020\|EPI_ISL_464873\|2020 | ···················· | ···················· | ···················· | ···················· | NNNNNNNNNNNNNNNNNNNN | NNNNNNNNNNNNNNNNNNNN | 1 |

| | | | | | | | | | |
|---|---|---|---|---|---|---|---|---|---|
| D | 7748 | hCoV-19/England/20118000304/2020\|EPI_ISL_464876\|20 | ···················· | ···················· | ···················· | ···················· | ·NNNNNNNNNNN········ | ··NN················ | 1 |
| D | 7749 | hCoV-19/England/201180048/2020\|EPI_ISL_464877\|2020 | ···················· | ···················· | ···················· | ···················· | NNNNNNNNNNNNNNNNNNNN | NNNNNNNNNNNNNNNNNNNN | 1 |
| D | 7752 | hCoV-19/England/201180095/2020\|EPI_ISL_464882\|2020 | ···················· | ···················· | ···················· | ···················· | ··NNNNNNNNNNNNNNNNNN | ·········N··NNNNNNNN | 1 |
| D | 7753 | hCoV-19/England/201180107/2020\|EPI_ISL_464883\|2020 | ···················· | ···················· | ···················· | ···················· | NNNNNNNNNNNNNNNNNNNN | NNNNNNNNNNNNNNNNNNNN | 1 |
| D | 7755 | hCoV-19/Switzerland/BS-ETHZ-120029/2020\|EPI_ISL_46 | ···················· | ···················· | ···················· | ···················· | ·······NNNNNNNNNNNNN | NNNNNNNNNNNNNNNNNNNN | 1 |
| D | 7758 | hCoV-19/Switzerland/BL-ETHZ-120033/2020\|EPI_ISL_46 | ···················· | ···················A | ···················· | ···················· | ·······NNNNNNNNNNNNN | NNNNNNNNNNNNNNNNNNNN | 1 |
| D | 7759 | hCoV-19/Switzerland/BL-ETHZ-120036/2020\|EPI_ISL_46 | ···················· | ···················· | ···················· | ···················· | ·······NNNNNNNNNNNNN | NNNNNNNNNNNNNNNNNNNN | 1 |
| D | 7760 | hCoV-19/Switzerland/BL-ETHZ-120038/2020\|EPI_ISL_46 | ···················· | ···················A | ···················· | ···················· | ·······NNNNNNN······ | ···················· | 1 |
| D | 7761 | hCoV-19/Switzerland/BL-ETHZ-120039/2020\|EPI_ISL_46 | ···················· | ···················· | ···················· | ···················· | ·······NNNNNNNNNNNNN | NNNNNNNNNNNNNNNNNNNN | 1 |
| D | 7763 | hCoV-19/Switzerland/BE-ETHZ-120050/2020\|EPI_ISL_46 | ···················· | ···················· | ···················· | ···Y················ | ·······NNNNNNNNNNNNN | NNNNNNNNNNNNNNNNNNNN | 1 |
| D | 8055 | hCoV-19/England/201161543/2020\|EPI_ISL_464855\|2020 | ···················· | ···················· | ···················· | ···················· | NNNNNNNNNNNNNNNNNNNN | ··N··N·NNNNNNNNNNNNN | 1 |
| D | 8056 | hCoV-19/England/201161565/2020\|EPI_ISL_464859\|2020 | ···················· | ···················· | ···················· | ···················· | ·NNNNNNNNNNNNNNNNNNN | NNNNNNNNNNNNNNNNNNNN | 1 |
| D | 8057 | hCoV-19/England/20116159902/2020\|EPI_ISL_464861\|20 | ···················· | ···················· | ···················· | ···················· | ·NNNNNNNNNNNNNNNNNNN | NNNNNNNNNNNNNNNNNNNN | 1 |
| D | 8061 | hCoV-19/England/201161635/2020\|EPI_ISL_464866\|2020 | ···················· | ···················· | ···················· | ···················· | NNNNNNNNNNNNNNNNNNNN | NNNNNNNNNNNNNNNNNNNN | 1 |
| D | 8062 | hCoV-19/England/201161656/2020\|EPI_ISL_464868\|2020 | ···················· | ···················· | ···················· | ···················· | ·NNNNNNNNNNNNNNNNNNN | NNNNNNNNN··N·NNNN··· | 1 |
| D | 8064 | hCoV-19/England/201161690/2020\|EPI_ISL_464871\|2020 | ···················· | ···················· | ···················· | ···················· | NNNNNNNNNNNNNNNNNNNN | NNNNNNNNNNNNNNNNNNNN | 1 |
| D | 8065 | hCoV-19/England/201180326/2020\|EPI_ISL_464897\|2020 | ···················· | ···················· | ···················· | ···················· | NNNNNNNNNNNNNNNNNNNN | NNNNNNNNNNNNNNNNNNNN | 1 |
| D | 8067 | hCoV-19/Spain/AN-IBV-001897/2020\|EPI_ISL_467071\|20 | ···················· | ···················· | ···················· | ···················· | NNNNNNNNNNNNNNNNNNNN | NNNNNNNNNNNNNNNNNNNN | 1 |
| D | 8068 | hCoV-19/Spain/AN-IBV-001899/2020\|EPI_ISL_467076\|20 | ···················· | ···················· | ···················· | ···················· | NNNNNNNNNNNNNNNNNNNN | NNNNNNNNNNNNNNNNNNNN | 1 |
| D | 8069 | hCoV-19/Spain/AN-IBV-001903/2020\|EPI_ISL_467077\|20 | ···················· | ···················· | ···················· | ···················· | NNNNNNNNNNNNNNNNNNNN | NNNNNNNNNNNNNNNNNNNN | 1 |
| D | 8070 | hCoV-19/Spain/AN-IBV-001915/2020\|EPI_ISL_467078\|20 | ···················· | ···················· | ···················· | ···················· | NNNNNNNNNNNNNNNNNNNN | NNNNNNNNNNNNNNNNNNNN | 1 |
| D | 8071 | hCoV-19/Spain/AN-IBV-001913/2020\|EPI_ISL_467079\|20 | ···················· | ···················· | ···················· | ···················· | NNNNNNNNNNNNNNNNNNNN | NNNNNNNNNNNNNNNNNNNN | 1 |
| D | 8072 | hCoV-19/Spain/AN-IBV-001919/2020\|EPI_ISL_467080\|20 | ···················· | ···················· | ···················· | ···················· | NNNNNNNNNNNNNNNNNNNN | NNNNNNNNNNNNNNNNNNNN | 1 |
| D | 8073 | hCoV-19/Spain/AN-IBV-001917/2020\|EPI_ISL_467081\|20 | ···················· | ···················· | ···················· | ···················· | NNNNNNNNNNNNNNNNNNNN | NNNNNNNNNNNNNNNNNNNN | 1 |
| D | 8074 | hCoV-19/Spain/AN-IBV-001905/2020\|EPI_ISL_467082\|20 | ···················· | ···················· | ···················· | ···················· | NNNNNNNNNNNNNNNNNNNN | NNNNNNNNNNNNNNNNNNNN | 1 |
| D | 8075 | hCoV-19/Spain/MD-IBV-001898/2020\|EPI_ISL_467083\|20 | ···················· | ···················· | ···················· | ···················· | NNNNNNNNNNNNNNNNNNNN | NNNNNNNNNNNNNNN····· | 1 |
| D | 8076 | hCoV-19/Spain/AN-IBV-001910/2020\|EPI_ISL_467084\|20 | ···················· | ···················· | ···················· | ···················· | NNNNNNNNNNNNNNNNNNNN | NNNNNNNNNNNNNNN····· | 1 |
| D | 8077 | hCoV-19/Spain/AN-IBV-001914/2020\|EPI_ISL_467085\|20 | ···················· | ···················· | ···················· | ···················· | NNNNNNNNNNNNNNNNNNNN | NNNNNNNNNNNNNNNNNNNN | 1 |
| D | 8079 | hCoV-19/Spain/CN-IBV-002600/2020\|EPI_ISL_467087\|20 | ···················· | ···················· | ···················· | ···················· | NNNNNNNNNNNNNNNNNNNN | NNNNNNNNNNNNNNNNNNNN | 1 |
| D | 8080 | hCoV-19/Spain/CN-IBV-002601/2020\|EPI_ISL_467088\|20 | ···················· | ···················· | ···················· | ···················· | NNNNNNNNNNNNNNNNNNNN | NNNNNNNNNNNNNNN····· | 1 |
| D | 8213 | hCoV-19/England/201160962/2020\|EPI_ISL_464810\|2020 | ···················· | ···················· | ···················· | ···················· | NNNNNNNNNNNNNNNNNNNN | ··N················· | 1 |
| D | 8214 | hCoV-19/England/201161147/2020\|EPI_ISL_464813\|2020 | ········A··········· | ···················· | ···················· | ···················· | ···················· | NNNN················ | 1 |
| D | 8215 | hCoV-19/England/201161151/2020\|EPI_ISL_464814\|2020 | ···················· | ···················· | ···················· | ···················· | ·NNNNNNNNNNNNNNNNNNN | ··N················· | 1 |
| D | 8216 | hCoV-19/England/201161176/2020\|EPI_ISL_464816\|2020 | ···················· | ···················· | ···················· | ···················· | NNNNNNNNNNNNNNNNNNNN | NNNNNNNNN·······N·N· | 1 |
| D | 8218 | hCoV-19/England/201161185/2020\|EPI_ISL_464818\|2020 | ···················· | ···················· | ···················· | ···················· | ·NNN······NNNN·N···· | ···················· | 1 |
| D | 8219 | hCoV-19/England/201161201/2020\|EPI_ISL_464820\|2020 | ···················· | ···················· | ···················· | ···················· | ··NNNNNNNNNNNNNNNN·· | ····N··············· | 1 |
| D | 8220 | hCoV-19/England/201161203/2020\|EPI_ISL_464821\|2020 | ···················· | ···················· | ···················· | ···················· | ··NNNNNNNNNNNNNN·NNN | NNNNNNNNNNNNNNNNNNNN | 1 |
| D | 8223 | hCoV-19/England/201161230/2020\|EPI_ISL_464826\|2020 | ···················· | ···················· | ···················· | ···················· | NNNNNNNNNNNNNNNNNNNN | NNNNNNNNNNNNNNNNNNNN | 1 |
| D | 8225 | hCoV-19/England/201161311/2020\|EPI_ISL_464831\|2020 | ···················· | ···················· | ···················· | ···················· | ··N····NN·N········· | ···················· | 1 |
| D | 8226 | hCoV-19/England/201161329/2020\|EPI_ISL_464833\|2020 | ···················· | ···················· | ···················· | ···················· | ···················· | NNNNN··NNNNNNNN····· | 1 |
| D | 8228 | hCoV-19/England/201161383/2020\|EPI_ISL_464836\|2020 | ···················· | ···················· | ···················· | ···················· | ···················· | NNNNNNNNNNNNNNNNNNNN | 1 |
| D | 8230 | hCoV-19/England/201161425/2020\|EPI_ISL_464839\|2020 | ···················· | ···················· | ···················· | ···················· | NNNNNNNNNNNNNNNNNNNN | NNNNNNNNNNNNNNNNNNNN | 1 |
| D | 8232 | hCoV-19/England/201161458/2020\|EPI_ISL_464842\|2020 | ···················· | ···················· | ···················· | ···················· | ···················· | NNNN················ | 1 |
| D | 8236 | hCoV-19/England/201161494/2020\|EPI_ISL_464850\|2020 | ···················· | ···················· | ···················· | ···················· | ·NNNNNNNNNNNNNNNNNNN | N··N················ | 1 |
| D | 8238 | hCoV-19/Switzerland/UR-ETHZ-130007/2020\|EPI_ISL_46 | ···················· | ···················· | ···················· | ···················· | ·······NNNNNNNNNNNNN | NNNNNNNNNNNNNNNNNNNN | 1 |
| D | 8240 | hCoV-19/Switzerland/BL-ETHZ-130012/2020\|EPI_ISL_46 | ···················· | ···················· | ···················· | ···················· | ·······NNNNNNNNNNNNN | NNNNNNNNNNNNNNNNNNNN | 1 |
| D | 8246 | hCoV-19/Spain/IB-IBV-002494/2020\|EPI_ISL_467055\|20 | ···················· | ···················· | ···················· | ···················· | NNNNNNNNNNNNNNNNNNNN | NNNNNNNNNNNNNNNNNNNN | 1 |
| D | 8247 | hCoV-19/Spain/IB-IBV-002492/2020\|EPI_ISL_467056\|20 | ···················· | ···················· | ···················· | ···················· | NNNNNNNNNNNNNNNNNNNN | NNNNNNNNNNNNNNNNNNNN | 1 |
| D | 8248 | hCoV-19/Spain/IB-IBV-002495/2020\|EPI_ISL_467058\|20 | ···················· | ···················· | ···················· | ···················· | NNNNNNNNNNNNNNNNNNNN | NNNNNNNNNNNNNNNNNNNN | 1 |
| D | 8249 | hCoV-19/Spain/AN-IBV-002897/2020\|EPI_ISL_467059\|20 | ···················· | ···················· | ···················· | ···················· | NNNNNNNNNNNNNNNNNNNN | NNNNNNNNNNNNNNNNNNNN | 1 |
| D | 8250 | hCoV-19/Spain/AN-IBV-002968/2020\|EPI_ISL_467061\|20 | ···················· | ···················· | ···················· | ···················· | NNNNNNNNNNNNNNNNNNNN | NNNNNNNNNNNNNNNNNNNN | 1 |
| D | 8252 | hCoV-19/Spain/AN-IBV-002987/2020\|EPI_ISL_467063\|20 | ···················· | ···················· | ···················· | ···················· | NNNNNNNNNNNNNNNNNNNN | NNNNNNNNNNNNNNNNNNNN | 1 |
| D | 8253 | hCoV-19/Spain/AN-IBV-001900/2020\|EPI_ISL_467064\|20 | ···················· | ···················· | ···················· | ···················· | NNNNNNNNNNNNNNNNNNNN | NNNNNNNNNNNNNNNNNNNN | 1 |
| D | 8254 | hCoV-19/Spain/MD-IBV-001911/2020\|EPI_ISL_467066\|20 | ···················· | ···················· | ···················· | ···················· | NNNNNNNNNNNNNNNNNNNN | NNNNNNNNNNNNNNNNNNNN | 1 |
| D | 8255 | hCoV-19/Spain/AN-IBV-001912/2020\|EPI_ISL_467068\|20 | ···················· | ···················· | ···················· | ···················· | NNNNNNNNNNNNNNNNNNNN | NNNNNNNNNNNNNNNNNNNN | 1 |
| D | 8256 | hCoV-19/Spain/AN-IBV-001920/2020\|EPI_ISL_467069\|20 | ···················· | ···················· | ···················· | ···················· | NNNNNNNNNNNNNNNNNNNN | NNNNNNNNNNNNNNNNNNNN | 1 |
| D | 8257 | hCoV-19/Spain/AN-IBV-001901/2020\|EPI_ISL_467070\|20 | ···················· | ···················· | ···················· | ···················· | NNNNNNNNNNNNNNNNNNNN | NNNNNNNNNNNNNNNNNNNN | 1 |
| D | 8258 | hCoV-19/Spain/AN-IBV-001907/2020\|EPI_ISL_467072\|20 | ···················· | ···················· | ···················· | ···················· | NNNNNNNNNNNNNNNNNNNN | NNNNNNNNNNNNNNNNNNNN | 1 |
| D | 8259 | hCoV-19/Spain/AN-IBV-001904/2020\|EPI_ISL_467073\|20 | ···················· | ···················· | ···················· | ···················· | NNNNNNNNNNNNNNNNNNNN | NNNNNNNNNNNNNNNNNNNN | 1 |
| D | 8260 | hCoV-19/Spain/AN-IBV-001902/2020\|EPI_ISL_467074\|20 | ···················· | ···················· | ···················· | ···················· | NNNNNNNNNNNNNNNNNNNN | NNNNNNNNNNNNNNNNNNNN | 1 |
| D | 8261 | hCoV-19/Spain/AN-IBV-001921/2020\|EPI_ISL_467075\|20 | ···················· | ···················· | ···················· | ···················· | NNNNNNNNNNNNNNNNNNNN | NNNNNNNNNNNNNNNNNNNN | 1 |
| D | 8263 | hCoV-19/Spain/CN-IBV-002592/2020\|EPI_ISL_467091\|20 | ···················· | ···················· | ···················· | ···················· | NNNNNNNNNNNNNNNNNNNN | ···················· | 1 |
| D | 8269 | hCoV-19/Spain/PV-IBV-001531/2020\|EPI_ISL_467097\|20 | ···················· | ···················· | ···················· | ···················· | NNNNNNN············· | ···················· | 1 |
| D | 8270 | hCoV-19/Spain/PV-IBV-001545/2020\|EPI_ISL_467098\|20 | ···················· | ···················· | ···················· | ···················· | NNNNNNNNNNNNNNNNNNNN | ···················· | 1 |
| D | 8354 | hCoV-19/England/EXET-1376D0/2020\|EPI_ISL_471982\|20 | ···················· | ···················· | ···················· | ···················· | NNNNNNNNNNNNNNNNNNNN | NNNNNNNNNNNNNNNNNNNN | 1 |
| D | 8355 | hCoV-19/England/EXET-137834/2020\|EPI_ISL_471987\|20 | ···················· | ···················· | ···················· | ···················· | NNNNNNNNNNNNNNNNNNNN | NNNNNNNNNNNNNNNNNNNN | 1 |
| D | 8358 | hCoV-19/England/EXET-1378BC/2020\|EPI_ISL_471991\|20 | ···················· | ···················· | ···················· | ···················· | NNNNNNNNNNNNNNNNNNNN | NNNNNNNNNNNNNNNNNNNN | 1 |
| D | 8508 | hCoV-19/England/20109053304/2020\|EPI_ISL_464583\|20 | ···················· | ···················· | ···················· | ···················· | ·NNNNNNNNNNNNNNNNNNN | ···················· | 1 |
| D | 8517 | hCoV-19/Switzerland/SH-ETHZ-130015/2020\|EPI_ISL_46 | ···················· | ···················· | ···················· | ···················· | ·······NNNNNNNNNNNNN | NNNNNNNNNNNNNNNNNNNN | 1 |
| D | 8518 | hCoV-19/Switzerland/BS-ETHZ-130020/2020\|EPI_ISL_46 | ···················· | ···················· | ···················· | ···················· | ·······NNNNNNNNNNNNN | NNNNNNNNNNNNNNNNNNNN | 1 |
| D | 8597 | hCoV-19/England/EXET-137139/2020\|EPI_ISL_471951\|20 | ···················· | ···················· | ···················· | ···················· | NNNNNNNNNNNNNNNNNNNN | NNNNNNNNNNNNNNNNNNNN | 1 |
| D | 8598 | hCoV-19/England/EXET-137218/2020\|EPI_ISL_471955\|20 | ···················· | ···················· | ···················· | ···················· | NNNNNNNNNNNNNNNNNNNN | NNNNNNNNNNNNNNNNNNNN | 1 |
| D | 8603 | hCoV-19/England/EXET-137324/2020\|EPI_ISL_471963\|20 | ···················· | ···················· | ···················· | ···················· | NNNNNNNNNNNNNNNNNNNN | NNNNNNNNNNNNNNNNNNNN | 1 |
| D | 8607 | hCoV-19/England/EXET-1373CA/2020\|EPI_ISL_471971\|20 | ···················· | ···················· | ···················· | ···················· | NNNNNNNNNNNNNNNNNNNN | NNNNNNNNNNNNNNNNNNNN | 1 |
| D | 8608 | hCoV-19/England/EXET-1373D9/2020\|EPI_ISL_471972\|20 | ···················· | ···················· | ···················· | ···················· | NNNNNNNNNNNNNNNNNNNN | NNNNNNNNNNNNNNNNNNNN | 1 |
| D | 8611 | hCoV-19/England/EXET-137412/2020\|EPI_ISL_471976\|20 | ···················· | ···················· | ···················· | ···················· | NNNNNNNNNNNNNNNNNNNN | NNNNNNNNNNNNNNNNNNNN | 1 |
| D | 8612 | hCoV-19/England/EXET-1374B8/2020\|EPI_ISL_471977\|20 | ···················· | ···················· | ···················· | ···················· | NNNNNNNNNNNNNNNNNNNN | NNNNNNNNNNNNNNNNNNNN | 1 |
| D | 8618 | hCoV-19/USA/NE-11554/2020\|EPI_ISL_475229\|2020-05-0 | ···················· | ···················· | ···················· | ···················· | NNNNNNNNNNNNNNNNNNNN | NNNNNNNNNNNNNNNNNNNN | 1 |
| D | 8630 | hCoV-19/England/PORT-2D0BE8/2020\|EPI_ISL_475263\|20 | ···················· | ···················· | ···················· | ···················· | NNNNNNNNNNNNNNNNNNNN | NNNNNNNNNNNNNNNNNNNN | 1 |
| D | 8746 | hCoV-19/England/20109002504/2020\|EPI_ISL_464505\|20 | ···················· | ···················· | ···················· | ···················· | ·NNNNNNNNNNNNNNNNNNN | ··NN·····N······N··· | 1 |

| | | | | | | | | | |
|---|---|---|---|---|---|---|---|---|---|
| D | 8747 | hCoV-19/England/20109002704/2020\|EPI_ISL_464506\|20 | ···················· | ···················· | ···················· | ···················· | ·NN·NNNNNNNNNNNNNNNN | NNNNNNNNNNNNNNNNNNNN | 1 |
| D | 8750 | hCoV-19/England/20109005404/2020\|EPI_ISL_464517\|20 | ···················· | ···················· | ···················· | ···················· | NNNNNNNNNNNNNNNNNNNN | NNNNNNNNNNNNNNNNNNNN | 1 |
| D | 8761 | hCoV-19/England/20109038б/2020\|EPI_ISL_464555\|2020 | ···················· | ···················· | ···················· | ···················· | ·NNNNNNNNNNNNNNNNNNN | NNNNNNNNNNNNNNNNNNNN | 1 |
| D | 8762 | hCoV-19/England/20109040104/2020\|EPI_ISL_464557\|20 | ···················· | ···················· | ···················· | ···················· | ··NNNNNNNNNNNNNNNNNN | ·········N·········· | 1 |
| D | 8763 | hCoV-19/England/20109041704/2020\|EPI_ISL_464559\|20 | ···················· | ···················· | ···················· | ···················· | ······N·NNN········· | ···················· | 1 |
| D | 8891 | hCoV-19/USA/NE-11505/2020\|EPI_ISL_475221\|2020-05-0 | ···················· | ···················· | ···················· | ···················· | NNNNNNNNNNNNNNNNNNNN | NNNNNNNNNNNNNNNNNNNN | 1 |
| D | 8973 | hCoV-19/England/20134058904/2020\|EPI_ISL_465811\|20 | ···················· | ···················· | ···················· | ···················· | ···················· | NNNN················ | 1 |
| D | 8974 | hCoV-19/England/20134075204/2020\|EPI_ISL_465812\|20 | ···················· | ···················· | ···················· | ···················· | ··NNNNNNNNNNNNNN···· | ···················· | 1 |
| D | 9180 | hCoV-19/Wales/PHWC-160771/2020\|EPI_ISL_472974\|2020 | ···················· | ···················· | ···················· | ···················· | NNNNNNNNNNNNNNNNNNNN | ···················· | 1 |
| D | 9198 | hCoV-19/Wales/PHWC-163C8E/2020\|EPI_ISL_474064\|2020 | ···················· | ···················· | ···················· | ···················· | NNNN················ | ···················· | 1 |
| D | 9206 | hCoV-19/USA/IA-10859/2020\|EPI_ISL_475190\|2020-05-0 | ···················· | ···················· | ···················· | ···················· | NNNNNNNNNNNNNNNNNNNN | NNNNNNNNNNNNNNNNNNNN | 1 |
| D | 9279 | hCoV-19/England/201100141/2020\|EPI_ISL_464643\|2020 | ···················· | ···················· | ···················· | ···················· | ···················· | NNNNNNNNNNNNNNN····· | 1 |
| D | 9280 | hCoV-19/England/201100347/2020\|EPI_ISL_464649\|2020 | ···················· | ···················· | ···················· | ···················· | ···················· | NNNNNNNNNNNNNNNNNNNN | 1 |
| D | 9282 | hCoV-19/England/201120015/2020\|EPI_ISL_464664\|2020 | ···················· | ···················· | ···················· | ···················· | ······NNNNNN·NNNNNNN | ···················· | 1 |
| D | 9293 | hCoV-19/England/20142139301/2020\|EPI_ISL_465966\|20 | ···················· | ···················· | ···················· | ···················· | ···NNNNNNNNNNNNNNNNN | NNNNNNNNNNNNNNNNNNNN | 1 |
| D | 9672 | hCoV-19/USA/MI-MDHHS-SC20684/2020\|EPI_ISL_471708\|2 | ···················· | ················A··· | ···················· | ···················· | NNN···········N····· | ···················· | 1 |
| E | 229 | hCoV-19/Romania/ROSV_4342/2020\|EPI_ISL_491074\|2020 | ···················· | ···················· | ···················· | ······NNNNNNNNNNNNNN | ···················· | ···················· | 1 |
| E | 232 | hCoV-19/Romania/ROSV_4666/2020\|EPI_ISL_491077\|2020 | ···················· | ···················· | ···················· | NNNNNNNNNNN········· | ···················· | ···················· | 1 |
| E | 238 | hCoV-19/Romania/ROSV_5230/2020\|EPI_ISL_491083\|2020 | ···················· | ···················· | ···················· | ·········NNNNNNNNNNN | ···················· | ···················· | 1 |
| E | 417 | hCoV-19/Romania/ROSV_11990/2020\|EPI_ISL_491039\|202 | ···················· | ···················· | ···················· | ·········C·GTAANNNN· | ···················· | ···················· | 1 |
| E | 418 | hCoV-19/Romania/ROSV_12235/2020\|EPI_ISL_491040\|202 | ···················· | ···················· | ···················· | ·········NNNNN······ | ···················· | ···················· | 1 |
| E | 419 | hCoV-19/Romania/ROSV_12278/2020\|EPI_ISL_491041\|202 | ···················· | ···················· | ···················· | ·········NNNNNNNNNNN | ···················· | ···················· | 1 |
| E | 422 | hCoV-19/Romania/ROSV_12587/2020\|EPI_ISL_491044\|202 | ···················· | ···················· | ···················· | ·········NNNNNN····· | ···················· | ···················· | 1 |
| E | 430 | hCoV-19/Romania/ROSV_1332/2020\|EPI_ISL_491052\|2020 | ···················· | ···················· | ···················· | ·········NNNNNN····· | ···················· | ···················· | 1 |
| E | 433 | hCoV-19/Romania/ROSV_1837/2020\|EPI_ISL_491055\|2020 | ···················· | ···················· | ···················· | ·············AANNNNN | ···················· | ···················· | 1 |
| E | 434 | hCoV-19/Romania/ROSV_1979/2020\|EPI_ISL_491056\|2020 | ···················· | ···················· | ···················· | ············TTNNNNNN | ···················· | ···················· | 1 |
| E | 435 | hCoV-19/Romania/ROSV_2113/2020\|EPI_ISL_491057\|2020 | ···················· | ···················· | ···················· | ·········NNNNNN····· | ···················· | ···················· | 1 |
| E | 437 | hCoV-19/Romania/ROSV_2778/2020\|EPI_ISL_491059\|2020 | ···················· | ···················· | ···················· | ············CTNNNNNN | ···················· | ···················· | 1 |
| E | 442 | hCoV-19/Romania/ROSV_5812/2020\|EPI_ISL_491086\|2020 | ···················· | ···················· | ···················· | ·········TNNNNNNNNNN | ···················· | ···················· | 1 |
| E | 445 | hCoV-19/Romania/ROSV_8235/2020\|EPI_ISL_491089\|2020 | ···················· | ···················· | ···················· | ·········A··NNNNNNNN | ···················· | ···················· | 1 |
| E | 447 | hCoV-19/Romania/ROSV_9826/2020\|EPI_ISL_491091\|2020 | ···················· | ···················· | ···················· | NNNNNNNNNNNNNN······ | ···················· | ···················· | 1 |
| E | 452 | hCoV-19/India/UN-11123/2020\|EPI_ISL_491096\|2020 | ···················· | ···················· | ···················· | ···················· | ····N····N·········· | NNN···N··N·N········ | 1 |
| E | 758 | hCoV-19/Scotland/CVR2451/2020\|EPI_ISL_493800\|2020- | ···················· | ···················A | ···················· | ···················· | ·····NNNNNNNNNNNNNNN | ···················· | 1 |
| E | 759 | hCoV-19/Scotland/CVR2464/2020\|EPI_ISL_493802\|2020- | ···················· | ···················· | ···················· | ···················· | NNNNNNNNNNNNNNNNNNNN | NNNNNNNNNNNNNNNNNNNN | 1 |
| E | 760 | hCoV-19/Scotland/CVR2541/2020\|EPI_ISL_493811\|2020- | ···················· | ···················A | ···················· | ···················· | NNNNNNNNNNNNNNNNNNNN | NNNNNNNNNNNNNNNNNNNN | 1 |
| E | 763 | hCoV-19/Scotland/CVR2613/2020\|EPI_ISL_493819\|2020- | ···················· | ···················A | ···················· | ···················· | NNNNNNNNNNNNNNNNNNNN | NNNNNNNNNNNNNNNNNNNN | 1 |
| E | 764 | hCoV-19/Scotland/CVR2626/2020\|EPI_ISL_493820\|2020- | ···················· | ···················A | ···················· | ···················· | NNNNNNNNNNNNNNNNNNNN | NNNNNNNNNNNNNNNNNNNN | 1 |
| E | 768 | hCoV-19/Scotland/CVR3261/2020\|EPI_ISL_493836\|2020- | ···················· | ·A·················· | ···················· | ···················· | ·······NNNNNNNNNNNNN | NNNNNNNNNNNNNNNNNNNN | 1 |
| E | 1009 | hCoV-19/Scotland/CVR2075/2020\|EPI_ISL_493762\|2020- | ···················· | ···················· | ···················· | ···················· | NNNNNNNNNNNNNNNNNNNN | NNNNNNNNNNNNNNNNNNNN | 1 |
| E | 1010 | hCoV-19/Scotland/CVR2077/2020\|EPI_ISL_493764\|2020- | ···T················ | ···················A | ···················· | ···················· | NNNNNNNNNNNNNNNNNNNN | NNNNNNNNNNNNNNNNNNNN | 1 |
| E | 1011 | hCoV-19/Scotland/CVR2097/2020\|EPI_ISL_493767\|2020- | ···················· | ···················· | ···················· | ···················· | NNNNNNNNNNNNNNNNNNNN | NNNNNNNNNNNNNNNNNNNN | 1 |
| E | 1012 | hCoV-19/Scotland/CVR2104/2020\|EPI_ISL_493769\|2020- | ···················· | ···················· | ···················· | ···················· | NNNNNNNNNNNNNNNNNNNN | NNNNNNNNNNNNNNNNNNNN | 1 |
| E | 1013 | hCoV-19/Scotland/CVR2105/2020\|EPI_ISL_493770\|2020- | ···················· | ···················· | ···················· | ···················· | NNNNNNNNNNNNNNNNNNNN | NNNNNNNNNNNNNNNNNNNN | 1 |
| E | 1014 | hCoV-19/Scotland/CVR2109/2020\|EPI_ISL_493771\|2020- | ···················· | ···················· | ···················· | ···················· | NNNNNNNNNNNNNNNNNNNN | ···················· | 1 |
| E | 1015 | hCoV-19/Scotland/CVR2111/2020\|EPI_ISL_493772\|2020- | ···················· | ···················· | ···················· | ···················· | NNNNNNNNNNNNNNNNNNNN | NNNNNNNNNNNNNNNNNNNN | 1 |
| E | 1016 | hCoV-19/Scotland/CVR2126/2020\|EPI_ISL_493773\|2020- | ···················· | ···················· | ···················· | ···················· | NNNNNNNNNNNNNNNNNNNN | NNNNNNNNNNNNNNNNNNNN | 1 |
| E | 1017 | hCoV-19/Scotland/CVR2137/2020\|EPI_ISL_493775\|2020- | ···················· | ···················· | ···················· | ···················· | NNNNNNNNNNNNNNNNNNNN | NNNNNNNNNNNNNNNNNNNN | 1 |
| E | 1018 | hCoV-19/Scotland/CVR2144/2020\|EPI_ISL_493777\|2020- | ···················· | ···················· | ···················· | ···················· | NNNNNNNNNNNNNNNNNNNN | NNNNNNNNNNNNNNNNNNNN | 1 |
| E | 1019 | hCoV-19/Scotland/CVR2155/2020\|EPI_ISL_493778\|2020- | ···················· | ·A·················· | ···················· | ···················· | NNNNNNNNNNNNNNNNNNNN | ···················· | 1 |
| E | 1020 | hCoV-19/Scotland/CVR2186/2020\|EPI_ISL_493782\|2020- | ···················· | ···················A | ···················· | ···················· | NNNNNNNNNNNNNNNNNNNN | ···················· | 1 |
| E | 1021 | hCoV-19/Scotland/CVR2188/2020\|EPI_ISL_493783\|2020- | ···················· | ···················· | ···················· | ···················· | NNNNNNNNNNNNNNNNNNNN | NNNNNNNNNNNNNNNNNNNN | 1 |
| E | 1022 | hCoV-19/Scotland/CVR2270/2020\|EPI_ISL_493785\|2020- | ···················· | ···················· | ···················· | ···················· | NNNNNNNNNNNNNNNNNNNN | ···················· | 1 |
| E | 1023 | hCoV-19/Scotland/CVR2336/2020\|EPI_ISL_493789\|2020- | ···················· | ···················· | ···················· | ···················· | NNNNNNNNNNNNNNNNNNNN | NNNNNNNNNNNNNNNNNNNN | 1 |
| E | 1024 | hCoV-19/Scotland/CVR2440/2020\|EPI_ISL_493799\|2020- | ···················· | ·A·················· | ···················· | ···················· | ·······NNNNNNNNNNNNN | ···················· | 1 |
| E | 1267 | hCoV-19/Scotland/CVR1909/2020\|EPI_ISL_493745\|2020- | ···················· | ···················· | ···················· | ···················· | ···················· | NNNNNNNNNNNNNNNNNNNN | 1 |
| E | 1268 | hCoV-19/Scotland/CVR1915/2020\|EPI_ISL_493746\|2020- | ···················· | ···················· | ···················· | ···················· | NNNNNNNNNNNNNNNNNNNN | NNNNNNNNNNNNNNNNNNNN | 1 |
| E | 1269 | hCoV-19/Scotland/CVR1916/2020\|EPI_ISL_493747\|2020- | ···················· | ···················A | ···················· | ···················· | NNNNNNNNNNNNNNNNNNNN | NNNNNNNNNNNNNNNNNNNN | 1 |
| E | 1270 | hCoV-19/Scotland/CVR1921/2020\|EPI_ISL_493748\|2020- | ···················· | ···················· | ···················· | ···················· | ·······NNNNNNNNNNNNN | ···················· | 1 |
| E | 1271 | hCoV-19/Scotland/CVR1955/2020\|EPI_ISL_493752\|2020- | ···················· | ···················· | ···················· | ···················· | NNNNNNNNNNNNNNNNNNNN | NNNNNNNNNNNNNNNNNNNN | 1 |
| E | 1272 | hCoV-19/Scotland/CVR1987/2020\|EPI_ISL_493753\|2020- | ···················· | ···················· | ···················· | ···················· | NNNNNNNNNNNNNNNNNNNN | NNNNNNNNNNNNNNNNNNNN | 1 |
| E | 1273 | hCoV-19/Scotland/CVR2019/2020\|EPI_ISL_493756\|2020- | ···················· | ···················· | ···················· | ···················· | NNNNNNNNNNNNNNNNNNNN | NNNNNNNNNNNNNNNNNNNN | 1 |
| E | 1274 | hCoV-19/Scotland/CVR2361/2020\|EPI_ISL_493790\|2020- | ···················· | ···················· | ···················· | ···················· | NNNNNNNNNNNNNNNNNNNN | ···················· | 1 |
| E | 1275 | hCoV-19/Scotland/CVR2364/2020\|EPI_ISL_493791\|2020- | ···················· | ···················· | ···················· | ···················· | NNNNNNNNNNNNNNNNNNNN | NNNNNNNNNNNNNNNNNNNN | 1 |
| E | 1641 | hCoV-19/Egypt/MASRI-007/2020\|EPI_ISL_482760\|2020-0 | ···················· | ···················· | ···················· | ·········C·NNNNNNNNN | ···················· | ···················· | 1 |
| E | 1642 | hCoV-19/Egypt/MASRI-008/2020\|EPI_ISL_482761\|2020-0 | ···················· | ···················· | ···················· | ·········NNNNNNNNNNN | ···················· | ···················· | 1 |
| E | 1643 | hCoV-19/Egypt/MASRI-011/2020\|EPI_ISL_482762\|2020-0 | ···················· | ···················· | ···················· | ············NNNN···· | ···················· | ···················· | 1 |
| E | 1646 | hCoV-19/Egypt/MASRI-014/2020\|EPI_ISL_482765\|2020-0 | ···················· | ···················· | ···················· | ·········NNNNNN····· | ···················· | ···················· | 1 |
| E | 1648 | hCoV-19/Egypt/MASRI-016/2020\|EPI_ISL_482767\|2020-0 | ········T··········· | ···················· | ···················· | NNNNNNNNNNNNNNNNNNNN | ···················· | ···················· | 1 |
| E | 1652 | hCoV-19/Egypt/MASRI-025/2020\|EPI_ISL_482771\|2020-0 | ···················· | ······A············· | ···················· | ·········ATG·NNNNNNN | ···················· | ···················· | 1 |
| E | 1670 | hCoV-19/Switzerland/VS-ETHZ-110053/2020\|EPI_ISL_48 | ···················· | ·A·················· | ···················· | ···················· | ·······NNNNNNNNNNNNN | ···················· | 1 |
| E | 1671 | hCoV-19/Switzerland/SG-ETHZ-110055/2020\|EPI_ISL_48 | ···················· | ···················· | ···················· | ···················· | ·······NNNNNNNNNNNNN | ···················· | 1 |
| E | 1700 | hCoV-19/Singapore/641/2020\|EPI_ISL_493401\|2020-04- | ···················· | ···················· | ···················· | ···················· | NNNNNNNNNNNNNNNNNNNN | ···················· | 1 |
| E | 1893 | hCoV-19/Switzerland/BS-ETHZ-110000/2020\|EPI_ISL_48 | R··················· | ···················· | ···················· | ···················· | ·······NNNNNNNNNNNNN | NNNNNNNNNNNNNNNNNNNN | 1 |
| E | 1894 | hCoV-19/Switzerland/BL-ETHZ-110003/2020\|EPI_ISL_48 | ···················· | ···················· | ···················· | ···················· | ·······NNNNNNNNNNNNN | NNNNNNNNNNNNNNNNNNNN | 1 |
| E | 1896 | hCoV-19/Switzerland/UR-ETHZ-110009/2020\|EPI_ISL_48 | ···················· | ···········R········ | ···················· | ···················· | ·······NNNNNNNNNNNNN | NNNNNNNNNNNNNNNNNNNN | 1 |
| E | 1897 | hCoV-19/Switzerland/VS-ETHZ-110010/2020\|EPI_ISL_48 | ···················· | ·A·················· | ···················· | ···················· | ·······NNNNNNNNNNNNN | NNNNNNNNNNNNNNNNNNNN | 1 |
| E | 1898 | hCoV-19/Switzerland/VS-ETHZ-110011/2020\|EPI_ISL_48 | ···················· | ···················· | ···················· | ···················· | ·······NNNNNNNNNNNNN | ···················· | 1 |
| E | 1899 | hCoV-19/Switzerland/VD-ETHZ-110013/2020\|EPI_ISL_48 | ···················· | ···················· | ···················· | ···················· | ·······NNNNNNNNNNNNN | NNNNNNNNNNNNNNNNNNNN | 1 |
| E | 1900 | hCoV-19/Switzerland/UR-ETHZ-110014/2020\|EPI_ISL_48 | ···················· | ···················· | ···················· | ···················· | ·······NNNNNNNNNNNNN | NNNNNNNNNNNNNNNNNNNN | 1 |
| E | 1901 | hCoV-19/Switzerland/BL-ETHZ-110015/2020\|EPI_ISL_48 | ···················· | ···················· | ···················· | ···················· | ·······NNNNNNNNNNNNN | NNNNNNNNNNNNNNNNNNNN | 1 |

| | | | | | | | | | |
|---|---|---|---|---|---|---|---|---|---|
| E | 1902 | hCoV-19/Switzerland/BS-ETHZ-110016/2020\|EPI_ISL_48 | ···················· | ···················· | ···················· | ···················· | ······NNNNNNNNNNNNNN | ···················· | 1 |
| E | 1903 | hCoV-19/Switzerland/BE-ETHZ-110017/2020\|EPI_ISL_48 | ···················· | W··················A | ···················· | ···················· | ······NNNNNNNNNNNNNN | ···················· | 1 |
| E | 1904 | hCoV-19/Switzerland/BE-ETHZ-110018/2020\|EPI_ISL_48 | ···················· | ···················· | ···················· | ···················· | ······NNNNNNNNNNNNNN | NNNNNNNNNNNNNNNNNNNN | 1 |
| E | 1905 | hCoV-19/Switzerland/BL-ETHZ-110020/2020\|EPI_ISL_48 | ···················· | ···················· | ···················· | ···················· | ······NNNNNNNNNNNNNN | NNNNNNNNNNNNNNNNNNNN | 1 |
| E | 1906 | hCoV-19/Switzerland/BL-ETHZ-110021/2020\|EPI_ISL_48 | ···················· | ···················· | ···················· | ···················· | ······NNNNNNNNNNNNNN | ···················· | 1 |
| E | 1907 | hCoV-19/Switzerland/VS-ETHZ-110024/2020\|EPI_ISL_48 | ···················· | ·A·················· | ···················· | ···················· | ······NNNNNNNNNNNNNN | NNNNNNNNNNNNNNNNNNNN | 1 |
| E | 1908 | hCoV-19/Switzerland/LU-ETHZ-110027/2020\|EPI_ISL_48 | ···················· | ·············M·M···· | ···················· | ···················· | ······NNNNNNNNNNNNNN | NNNNNNNNNNNNNNNNNNNN | 1 |
| E | 1909 | hCoV-19/Switzerland/LU-ETHZ-110029/2020\|EPI_ISL_48 | ···················· | ···················· | ···················· | ···················· | ······NNNNNNNNNNNNNN | ···················· | 1 |
| E | 1910 | hCoV-19/Switzerland/VS-ETHZ-110030/2020\|EPI_ISL_48 | ···················· | ···················· | ···················· | ···················· | ······NNNNNNNNNNNNNN | ···················· | 1 |
| E | 1911 | hCoV-19/Switzerland/BL-ETHZ-110034/2020\|EPI_ISL_48 | ···················· | ···················· | ···················· | ···················· | ······NNNNNNNNNNNNNN | ···················· | 1 |
| E | 1912 | hCoV-19/Switzerland/VD-ETHZ-110035/2020\|EPI_ISL_48 | ···················· | ···················· | ···················· | ···················· | ······NNNNNNNNNNNNNN | NNNNNNNNNNNNNNNNNNNN | 1 |
| E | 1913 | hCoV-19/Switzerland/BL-ETHZ-110036/2020\|EPI_ISL_48 | ···················· | ···················· | ···················· | ···················· | ······NNNNNNNNNNNNNN | NNNNNNNNNNNNNNNNNNNN | 1 |
| E | 1914 | hCoV-19/Switzerland/VS-ETHZ-110040/2020\|EPI_ISL_48 | ···················· | ···················· | ···················· | ···················· | ······NNNNNNNNNNNNNN | NNNNNNNNNNNNNNNNNNNN | 1 |
| E | 1915 | hCoV-19/Switzerland/VS-ETHZ-110042/2020\|EPI_ISL_48 | ···················· | ···················· | ···················· | ···················· | ······NNNNNNNNNNNNNN | NNNNNNNNNNNNNNNNNNNN | 1 |
| E | 1916 | hCoV-19/Switzerland/ZH-ETHZ-110046/2020\|EPI_ISL_48 | ···················· | ···················· | ···················· | ···················· | ······NNNNNNNNNNNNNN | NNNNNNNNNNNNNNNNNNNN | 1 |
| E | 1918 | hCoV-19/Switzerland/ZH-ETHZ-110050/2020\|EPI_ISL_48 | ···················· | ···················· | ···················· | ···················· | ······NNNNNNNNNNNNNN | NNNNNNNNNNNNNNNNNNNN | 1 |
| E | 1919 | hCoV-19/Switzerland/ZH-ETHZ-110058/2020\|EPI_ISL_48 | ···················· | ···················· | ···················· | ···················· | ······NNNNNNNNNNNNNN | ···················· | 1 |
| E | 1920 | hCoV-19/Switzerland/VD-ETHZ-110059/2020\|EPI_ISL_48 | ···················· | ···················· | ···················· | ···················· | ······NNNNNNNNNNNNNN | ···················· | 1 |
| E | 1921 | hCoV-19/Switzerland/BE-ETHZ-110060/2020\|EPI_ISL_48 | ···················· | ·A·················· | ···················· | ···················· | ······NNNNNNNNNNNNNN | NNNNNNNNNNNNNNNNNNNN | 1 |
| E | 1923 | hCoV-19/Switzerland/VD-ETHZ-110063/2020\|EPI_ISL_48 | ···················· | ···················· | ···················· | ···················· | ······NNNNNNNNNNNNNN | ···················· | 1 |
| E | 1924 | hCoV-19/Switzerland/AG-ETHZ-110065/2020\|EPI_ISL_48 | ···················· | ···················· | ···················· | ···················· | ······NNNNNNNNNNNNNN | NNNNNNNNNNNNNNNNNNNN | 1 |
| E | 1925 | hCoV-19/Switzerland/AG-ETHZ-110066/2020\|EPI_ISL_48 | ···················· | ···················· | ···················· | ···················· | ······NNNNNNNNNNNNNN | NNNNNNNNNNNNNNNNNNNN | 1 |
| E | 1926 | hCoV-19/Switzerland/SG-ETHZ-110067/2020\|EPI_ISL_48 | ···················· | ···················· | ···················· | ···················· | ······NNNNNNNNNNNNNN | NNNNNNNNNNNNNNNNNNNN | 1 |
| E | 1927 | hCoV-19/Switzerland/BE-ETHZ-110068/2020\|EPI_ISL_48 | ···················· | ·A·················· | ···················· | ···················· | ······NNNNNNNNNNNNNN | NNNNNNNNNNNNNNNNNNNN | 1 |
| E | 2284 | hCoV-19/England/NORW-EE81F/2020\|EPI_ISL_493542\|202 | ···················· | ···················· | ···················· | ···················· | NNNNNNNNNNNNNNNNNNNN | NNNNNNNNNNNNNNNNNNNN | 1 |
| E | 2285 | hCoV-19/England/NORW-EEB07/2020\|EPI_ISL_493543\|202 | ···················· | ···················· | ···················· | ···················· | NNNNNNNNNNNNNNNNNNNN | NNNNNNNNNNNNNNNNNNNN | 1 |
| E | 2286 | hCoV-19/England/NORW-EEB34/2020\|EPI_ISL_493544\|202 | ···················· | ···················· | ···················· | ···················· | NNNNNNNNNNNNNNNNNNNN | NNNNNNNNNNNNNNNNNNNN | 1 |
| E | 2384 | hCoV-19/USA/FL-BPHL-0168/2020\|EPI_ISL_480819\|2020- | ···················· | ···················· | ···················· | ···················· | NNNNNNNNN··········· | ···················· | 1 |
| E | 2595 | hCoV-19/USA/FL-BPHL-0151/2020\|EPI_ISL_480802\|2020- | ···················· | ···················· | ···················· | ···················· | NNNNNNNNNNNNNNNNNNNN | ···················· | 1 |
| E | 2596 | hCoV-19/USA/FL-BPHL-0153/2020\|EPI_ISL_480804\|2020- | ···················· | ···················· | ···················· | ···················· | NNNNNNNNNNNNNNNNNNNN | NNNNNNNNNNNNNNNNNNNN | 1 |
| E | 2597 | hCoV-19/USA/FL-BPHL-0154/2020\|EPI_ISL_480805\|2020- | ···················· | ···················· | ···················· | ···················· | NNNNNNNNNNNNNNNNNNNN | NNNNNNNNNNNNNNNNNNNN | 1 |
| E | 2600 | hCoV-19/USA/FL-BPHL-0158/2020\|EPI_ISL_480809\|2020- | ···················· | ···················· | ···················· | ···················· | NNNNNNNNNNNNNNNNNNNN | ···················· | 1 |
| E | 2602 | hCoV-19/USA/FL-BPHL-0184/2020\|EPI_ISL_480835\|2020- | ···················· | ···················· | ···················· | ···················· | NNNNNNNNNNNNNNNNNNNN | NNNNNNNNNNNNNNNNNNNN | 1 |
| E | 2611 | hCoV-19/USA/FL-BPHL-0195/2020\|EPI_ISL_480846\|2020- | ···················· | ···················· | ···················· | ···················· | NNNNNNNNNNNNNNNNNNNN | NNNNNNNNNNNNNNNNNNNN | 1 |
| E | 2744 | hCoV-19/India/UN-15348/2020\|EPI_ISL_491479\|2020 | ···················· | ···················· | ···················· | ···················· | NNNNNNNNNNNN········ | ···················· | 1 |
| E | 2840 | hCoV-19/Australia/VIC2161/2020\|EPI_ISL_480770\|2020 | ···················· | ···················· | ···················· | ···················· | NNNNNNNNNNNNNNNNNNNN | NNNNNNNNNNNNNNNNNNNN | 1 |
| E | 2843 | hCoV-19/Australia/VIC2166/2020\|EPI_ISL_480773\|2020 | ···················· | ···················· | ···················· | ···················· | NNNNNNNNNNNNNNNNNNNN | NNNNNNNNNNNNNNNNNNNN | 1 |
| E | 2844 | hCoV-19/Australia/VIC2167/2020\|EPI_ISL_480774\|2020 | ···················· | ···················· | ···················· | ···················· | ···················· | NNNNNNNNNNNNNNN····· | 1 |
| E | 2846 | hCoV-19/Australia/VIC2152/2020\|EPI_ISL_480776\|2020 | ···················· | ···················· | ···················· | ···················· | NNN···R··NNNNNNNNNNN | NNNNNNNNNNNNNNNNNNNN | 1 |
| E | 2847 | hCoV-19/Australia/VIC2162/2020\|EPI_ISL_480777\|2020 | ···················· | ···················· | ···················· | ···················· | NNNNNNNNNNNNNNNNNNNN | ···················· | 1 |
| E | 2859 | hCoV-19/Spain/PV-IBV-003215/2020\|EPI_ISL_480970\|20 | ···················· | ···················· | ···················· | ···················· | NNNNNNNNNNNNNNNNNNNN | NNNNNNNNNNNNNNNNNNNN | 1 |
| E | 2860 | hCoV-19/Spain/PV-IBV-003231/2020\|EPI_ISL_480971\|20 | ···················· | ···················· | ···················· | ···················· | NNNNNNNNNNNNNNNNNNNN | NNNNNNNNNNNNNNNNNNNN | 1 |
| E | 2861 | hCoV-19/Spain/PV-IBV-003200/2020\|EPI_ISL_480972\|20 | ···················· | ···················· | ···················· | ···················· | NNNNNNNNNNNNNNNNNNNN | NNNNNNNNNNNNNNNNNNNN | 1 |
| E | 2862 | hCoV-19/Spain/PV-IBV-003250/2020\|EPI_ISL_480973\|20 | ···················· | ···················· | ···················· | ···················· | NNNNNNNNNNNNNNNNNNNN | NNNNNNNNNNNNNNNNNNNN | 1 |
| E | 2863 | hCoV-19/Spain/CT-IBV-003738/2020\|EPI_ISL_480975\|20 | ···················· | ···················· | ···················· | ···················· | ··NNNNN············· | ···················· | 1 |
| E | 2864 | hCoV-19/Spain/PV-IBV-003246/2020\|EPI_ISL_480976\|20 | ···················· | ···················· | ···················· | ···················· | ·NNNNNNNNNNNNNNNNNNN | NNNNNNNNNNNNNNN····· | 1 |
| E | 2865 | hCoV-19/Spain/PV-IBV-003266/2020\|EPI_ISL_480977\|20 | ···················· | ···················· | ···················· | ···················· | NNNNNNNNNNNNNNNNNNNN | NNNNNNN·······N·NNNN | 1 |
| E | 2866 | hCoV-19/Spain/PV-IBV-003237/2020\|EPI_ISL_480978\|20 | ···················· | ···················· | ···················· | ···················· | NNNNNNNNNNNNNNNNNNNN | NNNNNNNNNNNNNNNNNNNN | 1 |
| E | 2867 | hCoV-19/Spain/PV-IBV-003210/2020\|EPI_ISL_480980\|20 | ···················· | ···················· | ···················· | ···················· | NNNNNNNNNNNNNNNNNNNN | N····NNNNNNNNNN····· | 1 |
| E | 2868 | hCoV-19/Spain/CT-IBV-003729/2020\|EPI_ISL_480981\|20 | ···················· | ···················· | ···················· | ···················· | NNNNNNNNNNNNNNNNNNNN | NNNNNNNNNNNNNNN····· | 1 |
| E | 2869 | hCoV-19/Spain/PV-IBV-003196/2020\|EPI_ISL_480982\|20 | ···················· | ···················· | ···················· | ···················· | NNNNNNNNNNNNNNNNNNNN | NNNNNNNNNNNNNNNNNNNN | 1 |
| E | 2870 | hCoV-19/Spain/PV-IBV-003238/2020\|EPI_ISL_480983\|20 | ···················· | ···················· | ···················· | ···················· | NNNNNNN·NNNNNNNNNNNN | NNN········NNNN····· | 1 |
| E | 2871 | hCoV-19/Spain/PV-IBV-003248/2020\|EPI_ISL_480984\|20 | ···················· | ···················· | ···················· | ···················· | NNNNNNNNNNNNNNNNNNNN | NNNNNNNNNNNNNNNNNNNN | 1 |
| E | 2872 | hCoV-19/Spain/PV-IBV-003228/2020\|EPI_ISL_480985\|20 | ···················· | ···················· | ···················· | ···················· | NNNNNNNNNNNNNNNNNNNN | NNNNNNNNNNNNNNNNNNNN | 1 |
| E | 2873 | hCoV-19/Spain/PV-IBV-003217/2020\|EPI_ISL_480986\|20 | ···················· | ···················· | ···················· | ···················· | NNNNNNNNNNNNNNNNNNNN | ···················· | 1 |
| E | 2874 | hCoV-19/Spain/PV-IBV-003249/2020\|EPI_ISL_480987\|20 | ···················· | ···················· | ···················· | ···················· | NNNNNNNNNNNNNNNNNNNN | NNNNNNNNNNNNNNNNNNNN | 1 |
| E | 2875 | hCoV-19/Spain/PV-IBV-003230/2020\|EPI_ISL_480988\|20 | ···················· | ···················· | ···················· | ···················· | NNNNNNNNNNNNNNNNNNNN | ···················· | 1 |
| E | 2876 | hCoV-19/Spain/CT-IBV-003726/2020\|EPI_ISL_480989\|20 | ···················· | ···················· | ···················· | ···················· | NNNNNNNNNNNNNNNNNNNN | NNNNNNNNNNNNNNNNNNNN | 1 |
| E | 3158 | hCoV-19/Spain/PV-IBV-003204/2020\|EPI_ISL_480952\|20 | ···················· | ···················· | ···················· | ···················· | NNNNNNNNNNNNNNNNNNNN | NNNNNNNNNNNNNNNNNNNN | 1 |
| E | 3159 | hCoV-19/Spain/PV-IBV-003257/2020\|EPI_ISL_480953\|20 | ···················· | ···················· | ···················· | ···················· | NNNNNNNNNNNNNNNNNNNN | NNNNNNNNNNNNNNNNNNNN | 1 |
| E | 3160 | hCoV-19/Spain/PV-IBV-003229/2020\|EPI_ISL_480954\|20 | ···················· | ···················· | ···················· | ···················· | NNNNNNNNNNNNNNNNNNNN | NNNNNNNNNNNNNNNNNNNN | 1 |
| E | 3161 | hCoV-19/Spain/PV-IBV-003263/2020\|EPI_ISL_480955\|20 | ···················· | ···················· | ···················· | ···················· | NNNNNNNNNNNNNNNNNNNN | NNNNNNNNNNNNNNN····· | 1 |
| E | 3162 | hCoV-19/Spain/PV-IBV-003226/2020\|EPI_ISL_480956\|20 | ···················· | ···················· | ···················· | ···················· | NNNNNNNNNNNNNNNNNNNN | NNNNNNNNNNNNNNNNNNNN | 1 |
| E | 3163 | hCoV-19/Spain/PV-IBV-003241/2020\|EPI_ISL_480957\|20 | ···················· | ···················· | ···················· | ···················· | NNNNNNNNNNNNNNNNNNNN | NNNNNNNNNNNNNNNNNNNN | 1 |
| E | 3164 | hCoV-19/Spain/PV-IBV-003259/2020\|EPI_ISL_480958\|20 | ···················· | ···················· | ···················· | ···················· | NNNNNNN············· | ···················· | 1 |
| E | 3166 | hCoV-19/Spain/PV-IBV-003203/2020\|EPI_ISL_480960\|20 | ···················· | ···················· | ···················· | ···················· | NNNNNNNNNNNNNNNNNNNN | NNNNNNNNNNNNNNNNNNNN | 1 |
| E | 3167 | hCoV-19/Spain/CT-IBV-003727/2020\|EPI_ISL_480961\|20 | ···················· | ···················· | ···················· | ···················· | NNNNNNNNNNNNNNNNNNNN | NNNNNNNNNNNNNNNNNNNN | 1 |
| E | 3168 | hCoV-19/Spain/PV-IBV-003190/2020\|EPI_ISL_480962\|20 | ···················· | ···················· | ···················· | ···················· | NNNNNNNNNNNNNNNNNNNN | ···········NNNN····· | 1 |
| E | 3169 | hCoV-19/Spain/PV-IBV-003221/2020\|EPI_ISL_480963\|20 | ···················· | ···················· | ···················· | ···················· | NNNNNNNNNNNNNNNNNNNN | NNNNNNNNNNNNNNNNNNNN | 1 |
| E | 3170 | hCoV-19/Spain/PV-IBV-003222/2020\|EPI_ISL_480965\|20 | ···················· | ···················· | ···················· | ···················· | NNNNNNNNNNN··NNNNNNN | NNNNNNNNNNNNNNN····· | 1 |
| E | 3171 | hCoV-19/Spain/PV-IBV-003251/2020\|EPI_ISL_480966\|20 | ···················· | ···················· | ···················· | ···················· | NNNNNNNNNNNNNNNNNNNN | NNNNNNNNNNNNNNNNNNNN | 1 |
| E | 3172 | hCoV-19/Spain/PV-IBV-003242/2020\|EPI_ISL_480967\|20 | ···················· | ···················· | ···················· | ···················· | NNNNNNNNNNNNNNNNNNNN | NNNNNNNNNNNNNNNNNNNN | 1 |
| E | 3173 | hCoV-19/Spain/PV-IBV-003223/2020\|EPI_ISL_480968\|20 | ···················· | ···················· | ···················· | ···················· | NNNNNNNNNNNNNNNNNNNN | NNNNNNNNNNNNNNNNNNNN | 1 |
| E | 3174 | hCoV-19/Spain/PV-IBV-003213/2020\|EPI_ISL_480969\|20 | ···················· | ···················· | ···················· | ···················· | NNNNNNNNNNNNNNNNNNNN | NNNNNNNNNNNNNNNNNNNN | 1 |
| E | 3175 | hCoV-19/Spain/PV-IBV-003234/2020\|EPI_ISL_480991\|20 | ···················· | ···················· | ···················· | ···················· | ·NNNNNNNNNNNNNNNNNNN | NNNNNNNNNNNNNNNNNNNN | 1 |
| E | 3176 | hCoV-19/Spain/PV-IBV-003256/2020\|EPI_ISL_480994\|20 | ···················· | ···················· | ···················· | ···················· | NNNNNNNNNNNNNNNNNNNN | NNNNNNNNNNNNNNN····· | 1 |
| E | 3177 | hCoV-19/Spain/CT-IBV-003733/2020\|EPI_ISL_480995\|20 | ···················· | ···················· | ···················· | ···················· | NNNNNNNNNNNNNNNNNNNN | NNNNNNNNNNNNNNNNNNNN | 1 |
| E | 3178 | hCoV-19/Spain/PV-IBV-003255/2020\|EPI_ISL_480996\|20 | ···················· | ···················· | ···················· | ···················· | NNNNNNNNNNNNNNNNNNNN | NNNNNNNNNNNNNNNNNNNN | 1 |
| E | 3179 | hCoV-19/Spain/PV-IBV-003265/2020\|EPI_ISL_480997\|20 | ···················· | ···················· | ···················· | ···················· | NNNNNNN············· | ···················· | 1 |
| E | 3180 | hCoV-19/Spain/PV-IBV-003225/2020\|EPI_ISL_480998\|20 | ···················· | ···················· | ···················· | ···················· | NNNNNNN············· | ···················· | 1 |

| | | | | | | | | | |
|---|---|---|---|---|---|---|---|---|---|
| E | 3181 | hCoV-19/Spain/PV-IBV-003227/2020\|EPI_ISL_480999\|20 | ···················· | ···················· | ···················· | ···················· | NNNNNNNNNNNNNNNNNNNN | ···················· | 1 |
| E | 3326 | hCoV-19/India/TG-CCMB-M288/2020\|EPI_ISL_495197\|202 | ···················· | ···················· | ···················· | ···················· | ·······NNNNNNNNNNNNN | NNNNNNNNNNNNNNNNNNNN | 1 |
| E | 3483 | hCoV-19/England/202691844/2020\|EPI_ISL_490225\|2020 | ···················· | ···················· | ···················· | ···················· | NNNNNNN···N········· | ···················· | 1 |
| E | 3487 | hCoV-19/England/202822172/2020\|EPI_ISL_490229\|2020 | ···················· | ············M······· | ···················· | ···················· | ···················· | NNNN·N·············· | 1 |
| E | 3556 | hCoV-19/India/TG-CCMB-L1892/2020\|EPI_ISL_495168\|20 | ···················· | ···················· | ···················· | ···················· | ··············N····· | NNNNNNNNNNNNNNNNNNNN | 1 |
| E | 3564 | hCoV-19/India/TG-CCMB-L1965/2020\|EPI_ISL_495176\|20 | ···················· | ···················· | ···················· | ···················· | ·······NNNNNNNNNNNNN | NNNNNNNNNNNNNNNNNNNN | 1 |
| E | 3612 | hCoV-19/India/MH-NIV-25949/2020\|EPI_ISL_479657\|202 | ···················· | ···················· | ···················· | ········C·NNNNNN···· | ···················· | ···················· | 1 |
| E | 3613 | hCoV-19/Australia/VIC1800/2020\|EPI_ISL_480607\|2020 | ···················· | ···················· | ···················· | ···················· | NNNNNNNNNNNNNNNNNNNN | NNNNNNNNNNNNNNNNNNNN | 1 |
| E | 3614 | hCoV-19/Australia/VIC1796/2020\|EPI_ISL_480608\|2020 | ···················· | ···················· | ···················· | ···················· | NNNNNNNNNNNNNNNNNNNN | NNNNNNNNNNNNNNNNNNNN | 1 |
| E | 3616 | hCoV-19/Australia/VIC1798/2020\|EPI_ISL_480622\|2020 | ···················· | ···················· | ···················· | ···················· | NNNNNNNNNNNNNNNNNNNN | NNNNNNNNNNNNNNNNNNNN | 1 |
| E | 3617 | hCoV-19/Australia/VIC1801/2020\|EPI_ISL_480627\|2020 | ···················· | ···················· | ···················· | ···················· | NNNNNNNNNNNNNNNNNNNN | NNNNNNNNNNNNNNNNNNNN | 1 |
| E | 3618 | hCoV-19/Australia/VIC1804/2020\|EPI_ISL_480630\|2020 | ···················· | ···················· | ···················· | ···················· | NNNNNNNNNNNNNNNNNNNN | NNNNNNNNNNNNNNNNNNNN | 1 |
| E | 3619 | hCoV-19/Australia/VIC1805/2020\|EPI_ISL_480631\|2020 | ···················· | ···················· | ···················· | ···················· | NNNNNNNNNNNNNNNNNNNN | NNNNNNNNNNNNNNNNNNNN | 1 |
| E | 3620 | hCoV-19/Australia/VIC1806/2020\|EPI_ISL_480632\|2020 | ···················· | ···················· | ···················· | ···················· | NNNNNNNNNNNNNNNNNNNN | NNNNNNNNNNNNNNNNNNNN | 1 |
| E | 3621 | hCoV-19/Australia/VIC1812/2020\|EPI_ISL_480636\|2020 | ···················· | ···················· | ···················· | ···················· | NNNNNNNNNNNNNNNNNNNN | NNNNNNNNNNNNNNNNNNNN | 1 |
| E | 3622 | hCoV-19/Australia/VIC1816/2020\|EPI_ISL_480640\|2020 | ···················· | ···················· | ···················· | ···················· | NNNNNNNNNNNNNNNNNNNN | NNNNNNNNNNNNNNNNNNNN | 1 |
| E | 3624 | hCoV-19/Australia/VIC1888/2020\|EPI_ISL_480642\|2020 | ···················· | ···················· | ···················· | ···················· | NNNNNNNNNNNNNNNNNNNN | NNNNNNNNNNNNNNNNNNNN | 1 |
| E | 3625 | hCoV-19/Australia/VIC1890/2020\|EPI_ISL_480643\|2020 | ···················· | ···················· | ···················· | ···················· | NNNNNNNNNNNNNNNNNNNN | NNNNNNNNNNNNNNNNNNNN | 1 |
| E | 3626 | hCoV-19/Australia/VIC1898/2020\|EPI_ISL_480645\|2020 | ···················· | ···················· | ···················· | ···················· | NNNNNNNNNNNNNNNNNNNN | NNNNNNNNNNNNNNNNNNNN | 1 |
| E | 3860 | hCoV-19/Australia/VIC1849/2020\|EPI_ISL_480560\|2020 | ···················· | ···················· | ···················· | ···················· | NNNNNNNNNNNNNNNNNNNN | NNNNNNNNNNNNNNNNNNNN | 1 |
| E | 3861 | hCoV-19/Australia/VIC1852/2020\|EPI_ISL_480561\|2020 | ···················· | ···················· | ···················· | ···················· | NNNNNNNNNNNNNNNNNNNN | NNNNNNNNNNNNNNNNNNNN | 1 |
| E | 3862 | hCoV-19/Australia/VIC1882/2020\|EPI_ISL_480592\|2020 | ···················· | ···················· | ···················· | ···················· | NNNNNNNNNNNNNNNNNNNN | NNNNNNNNNNNNNNNNNNNN | 1 |
| E | 3863 | hCoV-19/Australia/VIC2132/2020\|EPI_ISL_480730\|2020 | ···················· | ···················· | ···················· | ···················· | NNNNNNNNNNNNNNNNNNNN | NNNNNNNNNNNNNNNNNNNN | 1 |
| E | 3865 | hCoV-19/Australia/VIC2134/2020\|EPI_ISL_480732\|2020 | ···················· | ···················· | ···················· | ···················· | NNNNNNNNNNNNNNNNNNNN | ···················· | 1 |
| E | 3868 | hCoV-19/Australia/VIC2126/2020\|EPI_ISL_480735\|2020 | ···················· | ···················· | ···················· | ···················· | NNNNNNNNNNNNNNNNNNNN | NNNNNNNNN··········· | 1 |
| E | 3870 | hCoV-19/Australia/VIC2136/2020\|EPI_ISL_480737\|2020 | ···················· | ···················· | ···················· | ···················· | NNNNNNNNNNNNNNNNNNNN | ·············N······ | 1 |
| E | 3872 | hCoV-19/Australia/VIC2138/2020\|EPI_ISL_480739\|2020 | ···················· | ···················· | ···················· | ···················· | NNNN················ | ···················· | 1 |
| E | 3873 | hCoV-19/Australia/VIC2139/2020\|EPI_ISL_480740\|2020 | ···················· | ···················· | ···················· | ···················· | NNNNNNNNNNNNNNNNNNNN | NNNNNNNNNNNNNNNNNNNN | 1 |
| E | 3875 | hCoV-19/Australia/VIC2141/2020\|EPI_ISL_480742\|2020 | ···················· | ·A·················· | ···················· | ···················· | NNNNNNNNNNNNNNNNNNNN | ···A················ | 1 |
| E | 3878 | hCoV-19/Australia/VIC2060/2020\|EPI_ISL_480746\|2020 | ···················· | ···················· | ···················· | ···················· | NNNNNNNNNNNNNNNNNNNN | NNNNNNNNNNNNNNNNNNNN | 1 |
| E | 3879 | hCoV-19/Australia/VIC2044/2020\|EPI_ISL_480748\|2020 | ···················· | ···················· | ···················· | ···················· | NNNNNNNNNNNNNNNNNNNN | NNNNNNNNN··········· | 1 |
| E | 3881 | hCoV-19/Australia/VIC2048/2020\|EPI_ISL_480752\|2020 | ···················· | ···················· | ···················· | ···················· | NNNNNNNNNNNNNNNNNNNN | NNNNNNNNNNNNNNNNNNNN | 1 |
| E | 3882 | hCoV-19/Australia/VIC2056/2020\|EPI_ISL_480753\|2020 | ···················· | ···················· | ···················· | ···················· | NNNNNNNNNNNNNNNNNNN· | ···················· | 1 |
| E | 3923 | hCoV-19/Portugal/IGC3853/2020\|EPI_ISL_491208\|2020- | ···················· | ···················· | ···················· | ···················· | NNNNNNNNNNNNNNNNNNNN | NNNNNNNNNNNNNNNNNNNN | 1 |
| E | 3924 | hCoV-19/Portugal/IGC3857/2020\|EPI_ISL_491209\|2020- | ···················· | ···················· | ···················· | ···················· | ···················· | NNNNNNNNNNNNNNNNNNNN | 1 |
| E | 3925 | hCoV-19/Portugal/IGC3859/2020\|EPI_ISL_491210\|2020- | ···················· | ···················· | ···················· | ···················· | NNNNNNNNNNNNNNNNNNNN | NNNNNNNNNNNNNNNNNNNN | 1 |
| E | 3927 | hCoV-19/Portugal/IGC3996/2020\|EPI_ISL_491217\|2020- | ···················· | ···················· | ···················· | ···················· | NNNNNNNNNNNNNNNNNNNN | NNNNNNNNNNNNNNNNNNNN | 1 |
| E | 3928 | hCoV-19/Portugal/IGC4000/2020\|EPI_ISL_491219\|2020- | ···················· | ···················· | ···················· | ···················· | NNNNNNNNNNNNNNNNNNNN | NNNNNNNNNNNNNNN····· | 1 |
| E | 3929 | hCoV-19/Portugal/IGC4016/2020\|EPI_ISL_491222\|2020- | ···················· | ···················· | ···················· | ···················· | NNNNNNNNNNNNNNNNNNNN | NNNNNNNNNNNNNNNNNNNN | 1 |
| E | 3934 | hCoV-19/Portugal/IGC4095/2020\|EPI_ISL_491228\|2020- | ···················· | ···················· | ···················· | ···················· | NNNNNNNNNNNNNNNNNNNN | ···················· | 1 |
| E | 3938 | hCoV-19/Portugal/IGC4106/2020\|EPI_ISL_491232\|2020- | ···················· | ···················· | ···················· | ···················· | NNNNNNNNNNNNNNNNNNNN | ···················· | 1 |
| E | 3939 | hCoV-19/Portugal/IGC4108/2020\|EPI_ISL_491233\|2020- | ···················· | ···················· | ···················· | ···················· | ···················· | NNNNNNNNNNNNNNN····· | 1 |
| E | 3940 | hCoV-19/Portugal/IGC4124/2020\|EPI_ISL_491235\|2020- | ···················· | ···················· | ···················· | ···················· | ···················· | NNNNNNNNNNNNNNNNNNNN | 1 |
| E | 3941 | hCoV-19/Portugal/IGC4228/2020\|EPI_ISL_491237\|2020- | ···················· | ···················· | ···················· | ···················· | NNNNNNNNNNNNNNNNNNNN | NNNNNNNNNNNNNNNNNNNN | 1 |
| E | 3943 | hCoV-19/Portugal/IGC4238/2020\|EPI_ISL_491239\|2020- | ···················· | ···················· | ···················· | ···················· | NNNNNNNNNNNNNNNNNNNN | NNNNNNNNNNNNNNNNNNNN | 1 |
| E | 3944 | hCoV-19/Portugal/IGC4245/2020\|EPI_ISL_491240\|2020- | ···················· | ···················· | ···················· | ···················· | ···················· | NNNNNNNNNNNNNNNNNNNN | 1 |
| E | 3945 | hCoV-19/Portugal/IGC4250/2020\|EPI_ISL_491241\|2020- | ···················· | ···················· | ···················· | ···················· | NNNNNNNNNNNNNNNNNNNN | NNNNNNNNNNNNNNNNNNNN | 1 |
| E | 3947 | hCoV-19/Portugal/IGC4260/2020\|EPI_ISL_491243\|2020- | ···················· | ···················· | ···················· | ···················· | NNNNNNNNNNNNNNNNNNNN | NNNNNNNNNNNNNNNNNNNN | 1 |
| E | 3949 | hCoV-19/Portugal/IGC4392/2020\|EPI_ISL_491245\|2020- | ···················· | ···················· | ···················· | ···················· | NNNNNNNNNNNNNNNNNNNN | NNNNNNNNNNNNNNNNNNNN | 1 |
| E | 3952 | hCoV-19/Portugal/IGC4424/2020\|EPI_ISL_491249\|2020- | ···················· | ···················· | ···················· | ···················· | NNNNNNNNNNNNNNNNNNNN | NNNNNNNNNNNNNNNNNNNN | 1 |
| E | 3954 | hCoV-19/Portugal/IGC4426/2020\|EPI_ISL_491251\|2020- | ···················· | ···················· | ···················· | ···················· | NNNNNNNNNNNNNNNNNNNN | NNNNNNNNNNNNNNNNNNNN | 1 |
| E | 3958 | hCoV-19/Portugal/IGC4468/2020\|EPI_ISL_491255\|2020- | ···················· | ···················· | ···················· | ···················· | NNNNNNNNNNNNNNNNNNNN | NNNNNNNNNNNNNNNNNNNN | 1 |
| E | 3959 | hCoV-19/Portugal/IGC4471/2020\|EPI_ISL_491256\|2020- | ···················· | ···················· | ···················· | ···················· | NNNNNNNNNNNNNNNNNNNN | NNNNNNNNNNNNNNNNNNNN | 1 |
| E | 3960 | hCoV-19/Portugal/IGC4476/2020\|EPI_ISL_491257\|2020- | ···················· | ···················· | ···················· | ···················· | NNNNNNNNNNNNNNNNNNNN | NNNNNNNNNNNNNNNNNNNN | 1 |
| E | 3964 | hCoV-19/Portugal/IGC4501/2020\|EPI_ISL_491261\|2020- | ···················· | ···················· | ···················· | ···················· | NNNNNNNNNNNNNNNNNNNN | NNNNNNNNNNNNNNNNNNNN | 1 |
| E | 3969 | hCoV-19/Portugal/R1203/2020\|EPI_ISL_491270\|2020-06 | ···················· | ···················· | ···················· | ···················· | NNNN···NNNNNNNNNNNNN | NNNNNNNNNNNNNNNNNNNN | 1 |
| E | 3970 | hCoV-19/Portugal/R1237/2020\|EPI_ISL_491271\|2020-06 | ···················· | ···················· | ···················· | ···················· | NNNNNNNNNNNNNNNNNNNN | NNNNNNNNNNNNNNNNNNNN | 1 |
| E | 3972 | hCoV-19/Portugal/R1294/2020\|EPI_ISL_491274\|2020-06 | ···················· | ···················· | ···················· | ···················· | ···················· | NNNNNNNNNNNNNNNNNNNN | 1 |
| E | 3974 | hCoV-19/Portugal/R473/2020\|EPI_ISL_491289\|2020-05- | ···················· | ···················· | ···················· | ···················· | NNNN················ | NNNNNNNNNNNNNNNNNNNN | 1 |
| E | 3975 | hCoV-19/Portugal/R486/2020\|EPI_ISL_491291\|2020-05- | ···················· | ···················· | ···················· | ···················· | NNNNNNNNNNNNNNNNNNNN | ···········NNNN····· | 1 |
| E | 3976 | hCoV-19/Portugal/R493/2020\|EPI_ISL_491292\|2020-05- | ···················· | ···················· | ···················· | ···················· | NNNNNNNNNNNNNNNNNNNN | NNNNNNNNNNNNNNNNNNNN | 1 |
| E | 4079 | hCoV-19/Scotland/CVR3271/2020\|EPI_ISL_493841\|2020- | ···················· | ···················A | ···················· | ···················· | ·······NNNNNNNNNNNNN | ···················· | 1 |
| E | 4081 | hCoV-19/Scotland/CVR3285/2020\|EPI_ISL_493849\|2020- | ···················· | ···················A | ···················· | ···················· | ·······NNNNNNNNNNNNN | ···················· | 1 |
| E | 4082 | hCoV-19/Scotland/CVR3289/2020\|EPI_ISL_493851\|2020- | ···················· | ···················· | ···················· | ···················· | ·······NNNNNNNNNNNNN | ···················· | 1 |
| E | 4086 | hCoV-19/Scotland/CVR3315/2020\|EPI_ISL_493868\|2020- | ···················· | ···················· | ···················· | ···················· | NNNNNNNNNNNNNNNNNNNN | ···················· | 1 |
| E | 4087 | hCoV-19/Scotland/CVR3321/2020\|EPI_ISL_493873\|2020- | ···················· | ···················A | ···················· | ···················· | ·······NNNNNNNNNNNNN | ···················· | 1 |
| E | 4092 | hCoV-19/Scotland/CVR3887/2020\|EPI_ISL_493897\|2020- | ···················· | ···················· | ···················· | ···················· | NNNNNNNNNNNNNNNNNNNN | NNNNNNNNNNNNNNNNNNNN | 1 |
| E | 4111 | hCoV-19/Australia/VIC2097/2020\|EPI_ISL_480706\|2020 | ···················· | ···················· | ···················· | ···················· | NNNNNNNNNNNNNNNNNNNN | ···················· | 1 |
| E | 4112 | hCoV-19/Australia/VIC2098/2020\|EPI_ISL_480707\|2020 | ···················· | ···················· | ···················· | ···················· | NNNNNNNNNNNNNNNNNNNN | NNNNNNNNNNNNNNNNNNNN | 1 |
| E | 4113 | hCoV-19/Australia/VIC2099/2020\|EPI_ISL_480708\|2020 | ···················· | ···················· | ···················· | ···················· | NNNNNNNNNNNNNNNNNNNN | NNNNNNNNNNNNNNNNNNNN | 1 |
| E | 4114 | hCoV-19/Australia/VIC2101/2020\|EPI_ISL_480709\|2020 | ···················· | ···················· | ···················· | ···················· | NNNN················ | ···················· | 1 |
| E | 4119 | hCoV-19/Australia/VIC2106/2020\|EPI_ISL_480714\|2020 | ···················· | ···················· | ···················· | ···················· | NNNNNNNNNNNNNNNNNNNN | ···················· | 1 |
| E | 4120 | hCoV-19/Australia/VIC2108/2020\|EPI_ISL_480715\|2020 | ···················· | ···················· | ···················· | ···················· | NNNNNNNNNNNNNNNNNNNN | ···················· | 1 |
| E | 4121 | hCoV-19/Australia/VIC2110/2020\|EPI_ISL_480716\|2020 | ···················· | ···················· | ···················· | ···················· | NNNNNNNNNNNN········ | ···················· | 1 |
| E | 4126 | hCoV-19/Australia/VIC2117/2020\|EPI_ISL_480722\|2020 | ···················· | ···················· | ···················· | ···················· | NNNNNNNNNNNNNNNNN··· | R··················· | 1 |
| E | 4128 | hCoV-19/Australia/VIC2121/2020\|EPI_ISL_480725\|2020 | ···················· | ···················· | ···················· | ···················· | NNNNNNNNNNNNNNNNNNNN | ···················· | 1 |
| E | 4129 | hCoV-19/Australia/VIC2123/2020\|EPI_ISL_480726\|2020 | ···················· | ···················· | ···················· | ···················· | NNNNNNNNNNNNNNNNNNN· | NNNN···NNNNNNNN····· | 1 |
| E | 4130 | hCoV-19/Australia/VIC2124/2020\|EPI_ISL_480727\|2020 | ···················· | ···················· | ···················· | ···················· | NNNNNNNNNNNNNNNNNNNN | NNNNNNNNNNNNNNNNNNNN | 1 |
| E | 4134 | hCoV-19/Australia/VIC2050/2020\|EPI_ISL_480754\|2020 | ···················· | ···················· | ···················· | ···················· | NNNNNNNNNNNNNNNNNNNN | NNNNNNNNNNNNNNNNNNNN | 1 |
| E | 4142 | hCoV-19/Australia/VIC2159/2020\|EPI_ISL_480762\|2020 | ···················· | ···················· | ···················· | ···················· | NNNNNNNNNNNNNNNNNNNN | NNNNNNNNNNNNNNNNNNNN | 1 |

```
E   4143 hCoV-19/Australia/VIC2154/2020|EPI_ISL_480763|2020    ····················  ····················  ····················  ····················  NNNNNNNNNNNNNNNNNNNN  ····················  1
E   4144 hCoV-19/Australia/VIC2149/2020|EPI_ISL_480764|2020    ····················  ····················  ····················  ····················  NNNNNNNNNNNNNNNNN···  NNNNNNNNNNNNNNNNNNNN  1
E   4146 hCoV-19/Australia/VIC2150/2020|EPI_ISL_480766|2020    ····················  ····················  ····················  ····················  NNNNNNNNNNNNNNNNNNNN  NNNNNNNNNNNNNNNNNNNN  1
E   4147 hCoV-19/Australia/VIC2155/2020|EPI_ISL_480767|2020    ····················  ····················  ····················  ····················  NNNNNNNNNNNNNNNNNNNN  NNNNNNNNNNNNNNNNNNNN  1
E   4148 hCoV-19/Australia/VIC2157/2020|EPI_ISL_480768|2020    ····················  ····················  ····················  ····················  NNNNNNNNNNNNNNNNNNNN  NNNNNNNNNNNNNNNNNNNN  1
E   4301 hCoV-19/Singapore/629/2020|EPI_ISL_490079|2020-06-    ····················  ····················  ····················  ····················  ····················  NNNNNNNNNNNN········  1
E   4320 hCoV-19/USA/WI-UW-716/2020|EPI_ISL_491392|2020-06-    ····················  ····················  ····················  ····················  NNNNNNNNNNNNNNNNNNNN  NNNNNNNNNNNNNNNNNNNN  1
E   4353 hCoV-19/Australia/VIC1894/2020|EPI_ISL_480650|2020    ····················  ····················  ····················  ····················  NNNNNNNNNNNNNNNNNNNN  NNNNNNNNNNNNNNNNNNNN  1
E   4354 hCoV-19/Australia/VIC1896/2020|EPI_ISL_480652|2020    ····················  ····················  ····················  ····················  NNNNNNNNNNNNNNNNNNNN  NNNNNNNNNNNNNNNNNNNN  1
E   4355 hCoV-19/Australia/VIC1899/2020|EPI_ISL_480653|2020    ····················  ····················  ····················  ····················  NNNNNNNNNNNNNNNNNNNN  NNNNNNNNNNNNNNNNNNNN  1
E   4356 hCoV-19/Australia/VIC2012/2020|EPI_ISL_480654|2020    ····················  ····················  ····················  ····················  NNNN················  ····················  1
E   4357 hCoV-19/Australia/VIC2013/2020|EPI_ISL_480655|2020    ····················  ····················  ····················  ····················  NNNNNNNNNNNNNNNNNNNN  NNNNNNNNNNNNNNNNNNNN  1
E   4359 hCoV-19/Australia/VIC2018/2020|EPI_ISL_480660|2020    ····················  ····················  ····················  ····················  NNNNNN··············  ····················  1
E   4360 hCoV-19/Australia/VIC2019/2020|EPI_ISL_480661|2020    ····················  ····················  ····················  ····················  NNNNNNNNNNNNNNNNNNNN  NNNNNNNNNNNNNNNNN···  1
E   4364 hCoV-19/Australia/VIC2026/2020|EPI_ISL_480667|2020    ····················  ····················  ····················  ····················  NNNNNNNNNNNNNNNNNNNN  NNNNNNNNNNNNNNNNNNNN  1
E   4365 hCoV-19/Australia/VIC2027/2020|EPI_ISL_480668|2020    ····················  ····················  ····················  ····················  NNNN················  ····················  1
E   4368 hCoV-19/Australia/NT48/2020|EPI_ISL_480695|2020        ····················  ····················  ····················  ····················  NNNN················  ····················  1
E   4371 hCoV-19/Australia/VIC2087/2020|EPI_ISL_480699|2020    ····················  ····················  ····················  ····················  NNNNNNNNNNNNNNNNNNNN  ····················  1
E   4601 hCoV-19/Wales/PHWC-160331/2020|EPI_ISL_493976|2020    ····················  ····················  ····················  ····················  NNNNNNNNNNNNNNNNNNNN  ····················  1
E   4606 hCoV-19/Wales/PHWC-1663E0/2020|EPI_ISL_493994|2020    ····················  ····················  ····················  ····················  NNNNNNNNNNNNNNNNNNNN  ····················  1
E   4607 hCoV-19/Wales/PHWC-16641A/2020|EPI_ISL_493996|2020    ····················  ····················  ····················  ····················  NNNNNNNNNNNNNNNNNNNN  NNNNNNNNNNNNNNNNNNNN  1
E   4608 hCoV-19/Wales/PHWC-166429/2020|EPI_ISL_493997|2020    ····················  ····················  ····················  ····················  NNNNNNNNNNNNNNNNNNNN  NNNNNNNNNNNNNNNNNNNN  1
E   4610 hCoV-19/Australia/VIC1949/2020|EPI_ISL_480673|2020    ····················  ····················  ····················  ····················  NNNN················  ····················  1
E   4613 hCoV-19/Australia/VIC1952/2020|EPI_ISL_480676|2020    ····················  ····················  ····················  ····················  NNNN················  ····················  1
E   4614 hCoV-19/Australia/VIC1953/2020|EPI_ISL_480677|2020    ····················  ····················  ····················  ····················  NNNN················  ····················  1
E   4616 hCoV-19/Australia/VIC1955/2020|EPI_ISL_480679|2020    ····················  ····················  ····················  ····················  NNNN················  ····················  1
E   4618 hCoV-19/Australia/VIC1957/2020|EPI_ISL_480681|2020    ····················  ····················  ····················  ····················  ····················  NNNN················  1
E   4620 hCoV-19/Australia/VIC1960/2020|EPI_ISL_480683|2020    ····················  ····················  ····················  ····················  NNNNNNNNNNNNNNNN····  ····················  1
E   4622 hCoV-19/Australia/VIC2040/2020|EPI_ISL_480687|2020    ····················  ····················  ····················  ····················  ····················  NNNNNNNNNNNNNNNNNNNN  1
E   4778 hCoV-19/Scotland/CVR2020/2020|EPI_ISL_490651|2020-    ····················  ···················A  ····················  ····················  NNNNNNNNNNNNNNNNNNNN  NNNNNNNNNNNNNNNNNNNN  1
E   4779 hCoV-19/Scotland/CVR2022/2020|EPI_ISL_490652|2020-    ····················  ····················  ····················  ····················  NNNNNNNNNNNNNNNNNNNN  NNNNNNNNNNNNNNNNNNNN  1
E   4782 hCoV-19/Scotland/CVR2073/2020|EPI_ISL_490659|2020-    ····················  ····················  ····················  ····················  NNNNNNNNNNNNNNNNNNNN  NNNNNNNNNNNNNNNNNNNN  1
E   4783 hCoV-19/Scotland/CVR2092/2020|EPI_ISL_490661|2020-    ····················  ····················  ····················  ····················  NNNNNNNNNNNNNNNNNNNN  NNNNNNNNNNNNNNNNNNNN  1
E   4787 hCoV-19/Scotland/CVR2122/2020|EPI_ISL_490666|2020-    ····················  ·A··················  ····················  ····················  NNNNNNNNNNNNNNNNNNNN  NNNNNNNNNNNNNNNNNNNN  1
E   4788 hCoV-19/Scotland/CVR2139/2020|EPI_ISL_490669|2020-    ····················  ····················  ····················  ····················  NNNNNNNNNNNNNNNNNNNN  NNNNNNNNNNNNNNNNNNNN  1
E   4791 hCoV-19/Scotland/CVR2289/2020|EPI_ISL_490680|2020-    ····················  ·A··················  ····················  ····················  NNNNNNNNNNNNNNNNNNNN  NNNNNNNNNNNNNNNNNNNN  1
E   4794 hCoV-19/Scotland/CVR2369/2020|EPI_ISL_490687|2020-    ····················  ····················  ····················  ····················  NNNNNNNNNNNNNNNNNNNN  ····················  1
E   4863 hCoV-19/Wales/PHWC-166447/2020|EPI_ISL_493998|2020    ····················  ····················  ····················  ····················  ·NNNNNNNN···········  ····················  1
E   4866 hCoV-19/England/HECH-2599E25/2020|EPI_ISL_482112|2    ····················  ····················  ····················  ····················  NNNNNNNNNNNNNNNNNNNN  ··················NN  1
E   4948 hCoV-19/USA/MI-UM-2006S000969/2020|EPI_ISL_484776|    ····················  ····················  ····················  ····················  NNNNNNNNNNNNNNNNNNNN  ····················  1
E   4949 hCoV-19/USA/MI-UM-2006S000976/2020|EPI_ISL_484779|    ····················  ····················  ····················  ····················  NNNNNNNNNNNNNNNNNNNN  ····················  1
E   4950 hCoV-19/USA/MI-UM-2006S000984/2020|EPI_ISL_484785|    ····················  ····················  ····················  ····················  NNNNNNNNNNNNNNNNNNNN  ····················  1
E   4952 hCoV-19/USA/MI-UM-2006S001016/2020|EPI_ISL_484792|    ····················  ····················  ····················  ····················  NNNNNNNNNNNNNNNNNNNN  ····················  1
E   5019 hCoV-19/Wales/PHWC-166CE5/2020|EPI_ISL_494103|2020    ····················  ····················  ····················  ····················  NNNNNNNNNNNNNNNNNNNN  ····················  1
E   5026 hCoV-19/Wales/PHWC-166DA6/2020|EPI_ISL_494113|2020    ····················  ····················  ····················  ····················  NNNNNNNNNNNNNNNNNNNN  NNNNNNNNNNNNNNNNNNNN  1
E   5029 hCoV-19/Wales/PHWC-166E94/2020|EPI_ISL_494124|2020    ····················  ····················  ····················  ····················  NNNNNNNNNNNNNNNNNNNN  NNNNNNNNNNNNNNNNNNNN  1
E   5032 hCoV-19/Wales/PHWC-166EFE/2020|EPI_ISL_494129|2020    ····················  ····················  ····················  ····················  NNNNNNNNNNNNNNNNNNNN  ····················  1
E   5038 hCoV-19/Wales/PHWC-166FDD/2020|EPI_ISL_494138|2020    ····················  ····················  ····················  ····················  NNNNNNNNNNNNNNNNNNNN  NNNNNNNNNNNNNNNNNNNN  1
E   5039 hCoV-19/Wales/PHWC-167006/2020|EPI_ISL_494140|2020    ····················  ····················  ····················  ····················  ··NNNNNNNNNNNNNNN···  ····················  1
E   5100 hCoV-19/USA/WI-UW-773/2020|EPI_ISL_495477|2020-07-    ····················  ····················  ····················  ····················  NNNNNNNNNNNNNNNNNNNN  NNNNNNNNNNNNNNNNNNNN  1
E   5101 hCoV-19/USA/WI-UW-775/2020|EPI_ISL_495479|2020-07-    ····················  ····················  ····················  ····················  NNNNNNNNNNNNNNNNNNNN  NNNNNNNNNNNNNNNNNNNN  1
E   5112 hCoV-19/USA/WI-UW-787/2020|EPI_ISL_495491|2020-07-    ····················  ····················  ····················  ····················  NNNNNNNNNNNNNNNNNNNN  NNNNNNNNNNNNNNNNNNNN  1
E   5187 hCoV-19/USA/MI-UM-2006S000864/2020|EPI_ISL_484719|    ····················  ····················  ····················  ····················  NNNNNNNNNNNNNNNNNNNN  ····················  1
E   5188 hCoV-19/USA/MI-UM-2006S000866/2020|EPI_ISL_484720|    ····················  ····················  ····················  ····················  NNNNNNNNNNNNNNNNNNNN  ····················  1
E   5189 hCoV-19/USA/MI-UM-2006S000867/2020|EPI_ISL_484721|    ····················  ····················  ····················  ····················  NNNNNNNNNNNNNNNNNNNN  ····················  1
E   5192 hCoV-19/USA/MI-UM-2006S000874/2020|EPI_ISL_484726|    ····················  ····················  ····················  ····················  ····················  NNNNNNNNNNNNNNNNNNNN  1
E   5194 hCoV-19/USA/MI-UM-2006S000882/2020|EPI_ISL_484732|    ····················  ····················  ····················  ····················  NNNNNNNNNNNNNNNNNNNN  ····················  1
E   5196 hCoV-19/USA/MI-UM-2006S000899/2020|EPI_ISL_484745|    ····················  ····················  ····················  ····················  NNNNNNNNNNNNNNNNNNNN  ····················  1
E   5198 hCoV-19/USA/MI-UM-2006S000916/2020|EPI_ISL_484754|    ····················  ····················  ····················  ····················  ·······NNNNNNNNNNNNN  ····················  1
E   5199 hCoV-19/USA/MI-UM-2006S000927/2020|EPI_ISL_484758|    ····················  ····················  ····················  ····················  NNNNNNNNNNNNNNNNNNNN  ····················  1
E   5200 hCoV-19/USA/MI-UM-2006S000930/2020|EPI_ISL_484759|    ····················  ····················  ····················  ····················  NNNNNNNNNNNNNNNNNNNN  ····················  1
E   5201 hCoV-19/USA/MI-UM-2006S000931/2020|EPI_ISL_484760|    ····················  ····················  ····················  ····················  NNNNNNNNNNNNNNNNNNNN  ····················  1
E   5202 hCoV-19/USA/MI-UM-2006S000932/2020|EPI_ISL_484761|    ····················  ····················  ····················  ····················  NNNNNNNNNNNNNNNNNNNN  ····················  1
E   5206 hCoV-19/USA/MI-UM-2006S000954/2020|EPI_ISL_484769|    ····················  ····················  ····················  ····················  NNNNNNNNNNNNNNNNNNNN  ····················  1
E   5348 hCoV-19/Wales/PHWC-166D88/2020|EPI_ISL_494111|2020    ····················  ····················  ····················  ····················  NNNNNNNNNNNNNNNNNNNN  NNNNNNNNNNNNNNNNNNNN  1
E   5352 hCoV-19/Wales/PHWC-166E49/2020|EPI_ISL_494120|2020    ····················  ····················  ····················  ····················  NNNNNNNNNNNNNNNNNNNN  NNNNNNNNNNNNNNNNNNNN  1
E   5357 hCoV-19/USA/FL-BPHL-0498/2020|EPI_ISL_495404|2020-    ····················  ····················  ····················  ····················  NNNNNNNNNNNNNNNNNNNN  NNNNNNNNNNNNNNNNNNNN  1
E   5501 hCoV-19/USA/FL-BPHL-0332/2020|EPI_ISL_489741|2020-    ····················  ····················  ····················  ····················  NNNNNNNNNNNNNNNNNNNN  ····················  1
E   5600 hCoV-19/South Africa/KRISP-0485/2020|EPI_ISL_49556    ····················  ····················  ····················  ····················  ··NNNNNNNNNNNNNNNNNN  ····················  1
E   5606 hCoV-19/USA/MI-UM-2006S001057/2020|EPI_ISL_495571|    ····················  ····················  ····················  ····················  NNNNNNNNNNNNNNNNNNNN  ····················  1
E   5611 hCoV-19/USA/MI-UM-2006S001095/2020|EPI_ISL_495578|    ····················  ····················  ····················  ····················  NNNNNNNNNNNNNNNNNNNN  ····················  1
E   5615 hCoV-19/USA/MI-UM-2006S001104/2020|EPI_ISL_495582|    ····················  ····················  ····················  ····················  NNNNNNNNNNNNNNNNNNNN  ····················  1
E   5617 hCoV-19/USA/MI-UM-2006S001112/2020|EPI_ISL_495584|    ····················  ····················  ····················  ····················  NNNNNNNNNNNNNNNNNNNN  ····················  1
E   5620 hCoV-19/USA/MI-UM-2006S001116/2020|EPI_ISL_495587|    ····················  ····················  ····················  ····················  NNNNNNNNNNNNNNNNNNNN  ····················  1
E   5622 hCoV-19/USA/MI-UM-2006S001118/2020|EPI_ISL_495589|    ····················  ····················  ····················  ····················  NNNNNNNNNNNNNNNNNNNN  ····················  1
E   5623 hCoV-19/USA/MI-UM-2006S001119/2020|EPI_ISL_495590|    ····················  ····················  ····················  ····················  NNNNNNNNNNNNNNNNNNNN  ····················  1
E   5629 hCoV-19/USA/MI-UM-2006S005728/2020|EPI_ISL_495596|    ····················  ····················  ····················  ····················  NNNNNNNNNNNNNNNNNNNN  ····················  1
E   5858 hCoV-19/Scotland/CVR2542/2020|EPI_ISL_490692|2020-    ····················  ····················  ····················  ····················  NNNNNNNNNNNNNNNNNNNN  NNNNNNNNNNNNNNNNNNNN  1
E   5861 hCoV-19/Scotland/CVR3900/2020|EPI_ISL_490696|2020-    ····················  ····················  ····················  ····················  NNNNNNNNNNNNNNNNNNNN  NNNNNNNNNNNNNNNNNNNN  1
E   5878 hCoV-19/South Africa/KRISP-0402/2020|EPI_ISL_49552    ····················  ····················  ····················  ····················  NNNNNNNNNNNNNNNNNNNN  ····················  1
```

| | | | | | | | | | |
|---|---|---|---|---|---|---|---|---|---|
| E | 6070 | hCoV-19/India/TG-CCMB-M697/2020\|EPI_ISL_495225\|202 | ···················· | ···················· | ···················· | ···················· | ·······NNNN···NN···· | ···················· | 1 |
| E | 6071 | hCoV-19/India/TG-CCMB-M7/2020\|EPI_ISL_495226\|2020- | ···················· | ···················· | ···················· | ···················· | ·······NNNNNNNNNNNNN | NNNNNNNNNNNNNNNNNNNN | 1 |
| E | 6180 | hCoV-19/Scotland/CVR1885/2020\|EPI_ISL_484522\|2020- | ···················· | ···················· | ···················· | ···················· | ·······NNNNNNNNNNNNN | ···················· | 1 |
| E | 6333 | hCoV-19/England/LIVE-A1784/2020\|EPI_ISL_490336\|202 | ···················· | ···················· | ···················· | ···················· | ·······NNNNNNNNNNNNN | ···················· | 1 |
| E | 6343 | hCoV-19/England/LIVE-A2000/2020\|EPI_ISL_490373\|202 | ···················· | ···················· | ···················· | ···················· | ·······NNNNNNNNNNNNN | NNNNNNNNNNNNNNNNNNNN | 1 |
| E | 6344 | hCoV-19/England/LIVE-A202E/2020\|EPI_ISL_490374\|202 | ···················· | ···················· | ···················· | ···················· | ·······NNNNNNNNNNNNN | ···················· | 1 |
| E | 6347 | hCoV-19/England/LIVE-A2316/2020\|EPI_ISL_490379\|202 | ···················· | ···················· | ···················· | ···················· | ·······NNNNNNNNNNNNN | ···················· | 1 |
| E | 6384 | hCoV-19/India/TG-CCMB-M446/2020\|EPI_ISL_495205\|202 | ···················· | ···················· | ···················· | ···················· | ····NNNNNNN···N···NN | ···················· | 1 |
| E | 6385 | hCoV-19/India/TG-CCMB-M451/2020\|EPI_ISL_495206\|202 | ···················· | ···················· | ···················· | ···················· | ······NNNNNNNNNNNNNN | ·················NNN | 1 |
| E | 6394 | hCoV-19/India/TG-CCMB-M58/2020\|EPI_ISL_495215\|2020 | ···················· | ···················· | ···················· | ···················· | ···················· | NNNNNNNNNNNNNNNNNNNN | 1 |
| E | 6395 | hCoV-19/India/TG-CCMB-M581/2020\|EPI_ISL_495216\|202 | ···················· | ···················· | ···················· | ···················· | ····NNNNNNNNNNNNNNNN | ···················· | 1 |
| E | 6398 | hCoV-19/India/TG-CCMB-M651/2020\|EPI_ISL_495219\|202 | ···················· | ···················· | ···················· | ···················· | ·······NNNNNNNNNNNNN | NNNNNNNNNNNNNNNNNNNN | 1 |
| E | 6559 | hCoV-19/England/NORW-ED86B/2020\|EPI_ISL_490502\|202 | ···················· | ···················· | ···················· | ···················· | NNNNNNNNNNNNNNNNNNNN | NNNNNNNNNNNNNNNNNNNN | 1 |
| E | 6560 | hCoV-19/England/NORW-ED8C5/2020\|EPI_ISL_490504\|202 | ···················· | ···················· | ···················· | ···················· | NNNNNNNNNNNNNNNNNNNN | NNNNNNNNNNNNNNNNNNNN | 1 |
| E | 6561 | hCoV-19/England/NORW-ED92C/2020\|EPI_ISL_490505\|202 | ···················· | ···················· | ···················· | ···················· | NNNNNNNNNNNNNNNNNNNN | NNNNNNNNNNNNNNNNNNNN | 1 |
| E | 6562 | hCoV-19/England/NORW-ED977/2020\|EPI_ISL_490506\|202 | ···················· | ···················· | ···················· | ···················· | NNNNNNNNNNNNNNNNNNNN | NNNNNNNNNNNNNNNNNNNN | 1 |
| E | 6563 | hCoV-19/England/NORW-ED995/2020\|EPI_ISL_490507\|202 | ···················· | ···················· | ···················· | ···················· | NNNNNNNNNNNNNNNNNNNN | NNNNNNNNNNNNNNNNNNNN | 1 |
| E | 6564 | hCoV-19/England/NORW-ED9C2/2020\|EPI_ISL_490509\|202 | ···················· | ···················· | ···················· | ···················· | NNNNNNNNNNNNNNNNNNNN | NNNNNNNNNNNNNNN····· | 1 |
| E | 6565 | hCoV-19/England/NORW-EDA29/2020\|EPI_ISL_490511\|202 | ···················· | ···················· | ···················· | ···················· | NNNNNNNNNNNNNNNNNNNN | NNNNNNNNNNNNNNNNNNNN | 1 |
| E | 6566 | hCoV-19/England/NORW-EDA56/2020\|EPI_ISL_490514\|202 | ···················· | ···················· | ···················· | ···················· | NNNNNNNNNNNNNNNNNNNN | NNNNNNNNNNNNNNNNNNNN | 1 |
| E | 6567 | hCoV-19/England/NORW-EDAB0/2020\|EPI_ISL_490515\|202 | ···················· | ···················· | ···················· | ···················· | NNNNNNNNNNNNNNNNNNNN | NNNNNNNNNNNNNNNNNNNN | 1 |
| E | 6570 | hCoV-19/England/NORW-EE2A8/2020\|EPI_ISL_490524\|202 | ···················· | ···················· | ···················· | ···················· | NNNNNNNNNNNNNNNNNNNN | NNNNNNNNNNNNNNNNNNNN | 1 |
| E | 6571 | hCoV-19/England/NORW-EE2C6/2020\|EPI_ISL_490526\|202 | ···················· | ···················· | ···················· | ···················· | NNNNNNNNNNNNNNNNNNNN | NNNNNNNNNNNNNNNNNNNN | 1 |
| E | 6574 | hCoV-19/England/NORW-EE34B/2020\|EPI_ISL_490530\|202 | ···················· | ···················· | ···················· | ···················· | NNNNNNNNNNNNNNNNNNNN | NNNNNNNNNNNNNNNNNNNN | 1 |
| E | 6575 | hCoV-19/England/NORW-EE369/2020\|EPI_ISL_490532\|202 | ···················· | ···················· | ···················· | ···················· | NNNNNNNNNNNNNNNNNNNN | NNNNNNNNNNNNNNNNNNNN | 1 |
| E | 6576 | hCoV-19/England/NORW-EE387/2020\|EPI_ISL_490533\|202 | ···················· | ···················· | ···················· | ···················· | NNNNNNNNNNNNNNNNNNNN | NNNNNNNNN··········· | 1 |
| E | 6577 | hCoV-19/England/NORW-EE3E1/2020\|EPI_ISL_490537\|202 | ···················· | ···················· | ···················· | ···················· | NNNNNNNNNNNNNNNNNNNN | NNNN················ | 1 |
| E | 6578 | hCoV-19/England/NORW-EE3F0/2020\|EPI_ISL_490538\|202 | ···················· | ···················· | ··A················· | ···················· | NNNN················ | ···················· | 1 |
| E | 6581 | hCoV-19/England/NORW-EE466/2020\|EPI_ISL_490543\|202 | ···················· | ···················· | ···················· | ···················· | NNNNNNNNNNNNNNNNNNNN | NNNNNNNNNNNNNNNNNNNN | 1 |
| E | 6582 | hCoV-19/England/NORW-EE4EE/2020\|EPI_ISL_490549\|202 | ···················· | ···················· | ···················· | ···················· | NNNNNNNNNNNNNNNNNNNN | ···················· | 1 |
| E | 6583 | hCoV-19/England/NORW-EE4FD/2020\|EPI_ISL_490550\|202 | ···················· | ···················· | ···················· | ···················· | NNNNNNNNNNNN········ | ···················· | 1 |
| E | 6584 | hCoV-19/England/NORW-EE527/2020\|EPI_ISL_490551\|202 | ···················· | ···················· | ···················· | ···················· | NNNN················ | ···················· | 1 |
| E | 6585 | hCoV-19/England/NORW-EE800/2020\|EPI_ISL_490556\|202 | ···················· | ···················· | ···················· | ···················· | NNNNNNNNNNNNNNNNNNNN | NNNNNNNNNNNNNNNNNNNN | 1 |
| E | 6586 | hCoV-19/England/NORW-EE9B2/2020\|EPI_ISL_490557\|202 | ···················· | ···················· | ···················· | ···················· | NNNNNNNNNNNNNNNNNNNN | NNNNNNNNNNNNNNNNNNNN | 1 |
| E | 6587 | hCoV-19/England/NORW-EE9C1/2020\|EPI_ISL_490558\|202 | ···················· | ···················· | ···················· | ···················· | NNNNNNNNNNNNNNNNNNNN | NNNNNNNNNNNNNNNNNNNN | 1 |
| E | 6612 | hCoV-19/Wales/PHWC-16692A/2020\|EPI_ISL_494061\|2020 | ···················· | ···················· | ···················· | ···················· | NNNNNNNNNNNNNNNNNNNN | NNNNN··············· | 1 |
| E | 6615 | hCoV-19/Wales/PHWC-166975/2020\|EPI_ISL_494064\|2020 | ···················· | ···················· | ···················· | ···················· | NNNNNNNNNNNNNNNNNNNN | NNNNNNNNNNNNNNNNNNNN | 1 |
| E | 6630 | hCoV-19/Wales/PHWC-166BE8/2020\|EPI_ISL_494092\|2020 | ···················· | ···················· | ···················· | ···················· | ···················· | NNNN················ | 1 |
| E | 6633 | hCoV-19/Wales/PHWC-166C12/2020\|EPI_ISL_494095\|2020 | ···················· | ···················· | ···················· | ···················· | NNNNNNNNNNNNNNNNNNNN | ···················· | 1 |
| E | 6634 | hCoV-19/Wales/PHWC-166C30/2020\|EPI_ISL_494096\|2020 | ···················· | ···················· | ···················· | ···················· | N·····NNNNNNNNNNN··· | ···················· | 1 |
| E | 6635 | hCoV-19/Wales/PHWC-166C5E/2020\|EPI_ISL_494098\|2020 | ···················· | ···················· | ···················· | ···················· | NNNNNNNNNNNNNNNNNNNN | NNNN················ | 1 |
| E | 6638 | hCoV-19/USA/FL-BPHL-0482/2020\|EPI_ISL_495388\|2020- | ···················· | ···················· | ···················· | ···················· | NNNNNNNNNNNNNNNNNNNN | NNNNNNNNNNNNNNNNNNNN | 1 |
| E | 6642 | hCoV-19/USA/FL-BPHL-0488/2020\|EPI_ISL_495394\|2020- | ···················· | ···················· | ···················· | ···················· | NNNNNNNNNNNNNNNNNNNN | NNNNNNNNNNNNNNNNNNNN | 1 |
| E | 6646 | hCoV-19/USA/FL-BPHL-0492/2020\|EPI_ISL_495398\|2020- | ···················· | ···················· | ···················· | ···················· | NNNNNNNNNNNNNNNNNNNN | NNNNNNNNNNNNNNNNNNNN | 1 |
| E | 6820 | hCoV-19/Wales/PHWC-166492/2020\|EPI_ISL_494002\|2020 | ···················· | ···················· | ···················· | ···················· | ·NNNNNNNNNNNNNNNNNNN | ···················· | 1 |
| E | 6828 | hCoV-19/Wales/PHWC-166526/2020\|EPI_ISL_494011\|2020 | ···················· | ···················· | ···················· | ···················· | NNNNNNNNNNNNNNNNNNNN | NNNNNNNNNNNNNNNNN··· | 1 |
| E | 6829 | hCoV-19/Wales/PHWC-166535/2020\|EPI_ISL_494012\|2020 | ···················· | ···················· | ···················· | ···················· | NNNNNNNNNNNNNNNNNNNN | NNNNNNNNNNNNNNNNNNNN | 1 |
| E | 6830 | hCoV-19/Wales/PHWC-166544/2020\|EPI_ISL_494013\|2020 | ···················· | ···················· | ···················· | ···················· | NNNNNN·············· | ···················· | 1 |
| E | 6831 | hCoV-19/Wales/PHWC-166553/2020\|EPI_ISL_494014\|2020 | ···················· | ···················· | ···················· | ···················· | ··N·····NNNNNNNNNNNN | ···················· | 1 |
| E | 6834 | hCoV-19/Wales/PHWC-16659F/2020\|EPI_ISL_494017\|2020 | ···················· | ···················· | ···················· | ···················· | ·NNNNNNNNNNN········ | ···················· | 1 |
| E | 6835 | hCoV-19/Wales/PHWC-1665CC/2020\|EPI_ISL_494020\|2020 | ···················· | ···················· | ···················· | ···················· | NNNNNNNNNNNN········ | ···················· | 1 |
| E | 6844 | hCoV-19/Wales/PHWC-166711/2020\|EPI_ISL_494032\|2020 | ···················· | ···················· | ···················· | ···················· | NNNNNNNNNNNNNNNNNNNN | ···················· | 1 |
| E | 6845 | hCoV-19/Wales/PHWC-16673F/2020\|EPI_ISL_494034\|2020 | ···················· | ···················· | ···················· | ···················· | NNNNNNNNNNNNNNNNNNNN | NNNNNNNNNNNNNNNNNNNN | 1 |
| E | 6848 | hCoV-19/Wales/PHWC-166799/2020\|EPI_ISL_494038\|2020 | ···················· | ···················· | ···················· | ···················· | ·NNNNNNNNNNN········ | ···················· | 1 |
| E | 6854 | hCoV-19/Wales/PHWC-16682D/2020\|EPI_ISL_494046\|2020 | ···················· | ···················· | ···················· | ···················· | NNNNNNNNNNNNNNNNNNNN | ···················· | 1 |
| E | 6868 | hCoV-19/Wales/PHWC-166B51/2020\|EPI_ISL_494086\|2020 | ···················· | ···················· | ···················· | ···················· | NNNNNNNNNNNNNNNNNNNN | NNNNNNNNNNNNNNNNNNNN | 1 |
| E | 6898 | hCoV-19/USA/FL-BPHL-0440/2020\|EPI_ISL_495346\|2020- | ···················· | ···················· | ···················· | ···················· | ···NNNNNNNNNNNNNNNNN | ···················· | 1 |
| E | 6900 | hCoV-19/USA/FL-BPHL-0443/2020\|EPI_ISL_495349\|2020- | ···················· | ···················· | ···················· | ···················· | NNNNNNNNNNNNNNNNNNNN | ···················· | 1 |
| E | 7161 | hCoV-19/England/NORW-ED801/2020\|EPI_ISL_490498\|202 | ···················· | ···················· | ··A················· | ···················· | NNNNNNNNNNNNNNNNNNNN | NNNNNNNNNNNNNNNNNNNN | 1 |
| E | 7432 | hCoV-19/Belgium/UGent-125/2020\|EPI_ISL_481222\|2020 | ···················· | ···················· | ···················· | ···················· | NNNNNNNNNNNNNNNNNNNN | NNNNNNNNNNNNNNNNNNNN | 1 |
| E | 7433 | hCoV-19/Belgium/UGent-128/2020\|EPI_ISL_481224\|2020 | ···················· | ···················· | ···················· | ···················· | NNNNNNNNNNNNNNNNNNNN | NNNNNNNNNNNNNNNNNNNN | 1 |
| E | 7436 | hCoV-19/Belgium/UGent-136/2020\|EPI_ISL_481229\|2020 | ···················· | ···················· | ···················· | ···················· | NNNNNNNNNNNNNNNNNNNN | NNNNNNNNNNNNNNNNNNNN | 1 |
| E | 7437 | hCoV-19/Belgium/UGent-138/2020\|EPI_ISL_481230\|2020 | ···················· | ···················· | ···················· | ···················· | NNNNNNNNNNNNNNNNNNNN | NNNNNNNNNNNNNNNNNNNN | 1 |
| E | 7439 | hCoV-19/Belgium/UGent-141/2020\|EPI_ISL_481232\|2020 | ···················· | ···················· | ···················· | ···················· | NNNNNNNNNNNNNNNNNNNN | NNNNNNNNNNNNNNNNNNNN | 1 |
| E | 8010 | hCoV-19/Singapore/6nan/2020\|EPI_ISL_482680\|2020-02 | ···················· | ···················· | ···················· | ···················· | ···················· | NN·N···NN··········· | 1 |
| E | 8012 | hCoV-19/Singapore/318/2020\|EPI_ISL_482683\|2020-06- | ···················· | ···················· | ···················· | ···················· | ···················· | NN·····NN··········· | 1 |
| E | 8013 | hCoV-19/Singapore/316/2020\|EPI_ISL_482684\|2020-06- | ···················· | ···················· | ···················· | ···················· | NNNNNNNNNNNNNNNNNNNN | NNNNNNNNNNNNNNNNNNNN | 1 |
| E | 8014 | hCoV-19/Singapore/537/2020\|EPI_ISL_482685\|2020-06- | ···················· | ···················· | ···················· | ···················· | NNNNNNNNNNNNNNNNNNNN | NNNNNNNNNNNNNNNNNNNN | 1 |
| E | 8015 | hCoV-19/Singapore/538/2020\|EPI_ISL_482686\|2020-06- | ···················· | ···················· | ···················· | ···················· | ··············N····· | NN·····NN··N········ | 1 |
| E | 8017 | hCoV-19/Singapore/540/2020\|EPI_ISL_482688\|2020-06- | ···················· | ···················· | ···················· | ···················· | ···N····NN····N····· | NNNNN··NNNNNN··N···· | 1 |
| E | 8018 | hCoV-19/Singapore/541/2020\|EPI_ISL_482689\|2020-06- | ···················· | ···················· | ···················· | ···················· | NNNNNNNNNNNNNNNNNNNN | NNNNNNNNNNNNNNNNNNNN | 1 |
| E | 8019 | hCoV-19/Singapore/542/2020\|EPI_ISL_482690\|2020-06- | ···················· | ···················· | ···················· | ···················· | NNNNNNNNNNNNNNNNNNNN | NNNNNNNNNNNNNNNNNNNN | 1 |
| E | 8020 | hCoV-19/Singapore/543/2020\|EPI_ISL_482691\|2020-06- | ···················· | ···················· | ···················· | ···················· | NNNNNNNNNNNNNNNNNNNN | NNNNNNNNNNNNNNNNNNNN | 1 |
| E | 8140 | hCoV-19/Mexico/GUA-InDRE-19/2020\|EPI_ISL_493334\|20 | ···················· | ···················· | ···················· | ···················· | NNNNNNNNNNNNNNNNNNNN | ···················· | 1 |
| E | 8157 | hCoV-19/Norway/2828/2020\|EPI_ISL_493381\|2020-06-29 | NNNNNNNNNNNNNNNNNNNN | ···················· | ···················· | ···················· | ···················· | ···················· | 1 |
| E | 8363 | hCoV-19/Mexico/BCS-InDRE_31/2020\|EPI_ISL_493346\|20 | ···················· | ·A·················· | ···················· | ···················· | ···················· | NNNNNNNNNNNNNNNNNNNN | 1 |
| E | 8449 | hCoV-19/Spain/PV-IBV-003244/2020\|EPI_ISL_481022\|20 | ···················· | ···················· | ···················· | ···················· | NNNNNNNNNNNNNNNNNNNN | NNNNNNNNNNNNNNNNNNNN | 1 |
| E | 8450 | hCoV-19/Spain/PV-IBV-003258/2020\|EPI_ISL_481023\|20 | ···················· | ···················· | ···················· | ···················· | NNNNNNN············· | ···················· | 1 |
| E | 8451 | hCoV-19/Spain/CT-IBV-003730/2020\|EPI_ISL_481025\|20 | ···················· | ···················· | ···················· | ···················· | NNNNNNNNNNNNNNNNNNNN | NNNNNNNNNNNNNNNNNNNN | 1 |
| E | 8452 | hCoV-19/Spain/PV-IBV-003192/2020\|EPI_ISL_481027\|20 | ···················· | ···················· | ···················· | ···················· | NNNNNNNNNNNNNNNNNNNN | NNNNNNNNNNNNNNNNNNNN | 1 |

| | | | | | | | | |
|---|---|---|---|---|---|---|---|---|
| E | 8453 hCoV-19/Spain/PV-IBV-003264/2020\|EPI_ISL_481028\|20 | ···················· | ···················· | ···················· | ···················· | NNNNNNNNNNNNNNNNNNNN | NNNNNNNNNNNNNNN··NNN | 1 |
| E | 8454 hCoV-19/Spain/CT-IBV-003736/2020\|EPI_ISL_481029\|20 | ···················· | ···················· | ···················· | ···················· | NNNNNNNNNNNNNNNNNNNN | NNNNNNNNNNNNNNNNNNNN | 1 |
| E | 8455 hCoV-19/Spain/PV-IBV-003212/2020\|EPI_ISL_481030\|20 | ···················· | ···················· | ···················· | ···················· | NNNNNNNNNNNNNNNNNNNN | NNNNNNNNNNNNNNNNNNNN | 1 |
| E | 8456 hCoV-19/Spain/MD-IBV-004930/2020\|EPI_ISL_481065\|20 | ···················· | ···················· | ···················· | ···················· | NNNNNNNNNNNNNNNNNNNN | NNNNNNNNNNNNNNNNNNNN | 1 |
| E | 8457 hCoV-19/Spain/MD-IBV-004914/2020\|EPI_ISL_481066\|20 | ···················· | ···················· | ···················· | ···················· | NNNNNNNNNNNNNNNNNNNN | NNNNNNNNNNNNNNNNNNNN | 1 |
| E | 8458 hCoV-19/Spain/MD-IBV-004979/2020\|EPI_ISL_481067\|20 | ···················· | ···················· | ···················· | ···················· | NNNNNNNNNNNNNNNNNNNN | NNNNNNNNNNNNNNNNNNNN | 1 |
| E | 8459 hCoV-19/Spain/MD-IBV-004973/2020\|EPI_ISL_481068\|20 | ···················· | ···················· | ···················· | ···················· | NNNNNNNNNNNNNNNNNNNN | ···················· | 1 |
| E | 8460 hCoV-19/Spain/MD-IBV-004952/2020\|EPI_ISL_481069\|20 | ···················· | ···················· | ···················· | ···················· | NNNNNNNNNNNNNNNNNNNN | NNNNNNNNNNNNNNNNNNNN | 1 |
| E | 8461 hCoV-19/Spain/MD-IBV-004949/2020\|EPI_ISL_481070\|20 | ···················· | ···················· | ···················· | ···················· | NNNNNNNNNNNNNNNNNNNN | NNNNNNNNNNNNNNNNNNNN | 1 |
| E | 8462 hCoV-19/Spain/MD-IBV-004955/2020\|EPI_ISL_481071\|20 | ···················· | ···················· | ···················· | ···················· | NNNNNNNNNNNNNNNNNNNN | NNNNNNNNNNNNNNNNNNNN | 1 |
| E | 8463 hCoV-19/Spain/MD-IBV-004931/2020\|EPI_ISL_481072\|20 | ···················· | ···················· | ···················· | ···················· | NNNNNNNNNNNNNNNNNNNN | NNNNNNNNNNNNNNNNNNNN | 1 |
| E | 8464 hCoV-19/Spain/MD-IBV-004971/2020\|EPI_ISL_481073\|20 | ···················· | ···················· | ···················· | ···················· | NNNNNNNNNNNNNNNNNNNN | NNNNNNNNNNNNNNN··NNN | 1 |
| E | 8465 hCoV-19/Spain/MD-IBV-004940/2020\|EPI_ISL_481074\|20 | ···················· | ···················· | ···················· | ···················· | NNNNNNN············· | ···················· | 1 |
| E | 8466 hCoV-19/Spain/MD-IBV-004898/2020\|EPI_ISL_481075\|20 | ···················· | ···················· | ···················· | ···················· | NNNNNNNNNNNNNNNNNNNN | NNNNNNNNNNNNNNN····· | 1 |
| E | 8467 hCoV-19/Spain/MD-IBV-004950/2020\|EPI_ISL_481076\|20 | ···················· | ···················· | ···················· | ···················· | NNNNNNNNNNNNNNNNNNNN | NNNNNNNNNNNNNNNNNNNN | 1 |
| E | 8468 hCoV-19/Spain/MD-IBV-004908/2020\|EPI_ISL_481077\|20 | ···················· | ···················· | ···················· | ···················· | ··NNNNN············· | ···················· | 1 |
| E | 8469 hCoV-19/Spain/MD-IBV-004935/2020\|EPI_ISL_481078\|20 | ···················· | ···················· | ···················· | ···················· | NNNNNNNNNNNNNNNNNNNN | NNNNNNNNNNNNNNNNNNNN | 1 |
| E | 8470 hCoV-19/Spain/MD-IBV-004959/2020\|EPI_ISL_481079\|20 | ···················· | ···················· | ···················· | ···················· | NNNNNNNNNNNNNNNNNNNN | NNNNNNNNNNNNNNNNNNNN | 1 |
| E | 8471 hCoV-19/Spain/MD-IBV-004928/2020\|EPI_ISL_481080\|20 | ···················· | ···················· | ···················· | ···················· | NNNNNNNNNNNNNNNNNNNN | NNNNNNNNNNNNNNNNNNNN | 1 |
| E | 8472 hCoV-19/Spain/MD-IBV-004939/2020\|EPI_ISL_481081\|20 | ···················· | ···················· | ···················· | ···················· | NNNNNNNNNNNNNNNNNNNN | NNNNNNNNNNNNNNNNNNNN | 1 |
| E | 8473 hCoV-19/Spain/MD-IBV-004951/2020\|EPI_ISL_481082\|20 | ···················· | ···················· | ···················· | ···················· | NNNNNNN··NN········· | NNNNNNNNNNNNNNN····· | 1 |
| E | 8474 hCoV-19/Spain/MD-IBV-004913/2020\|EPI_ISL_481083\|20 | ···················· | ···················· | ···················· | ···················· | NNNNNNNNNNNNNNNNNNNN | NNNNNNNNNNNNNNNNNNNN | 1 |
| E | 8475 hCoV-19/Spain/MD-IBV-004953/2020\|EPI_ISL_481084\|20 | ···················· | ···················· | ···················· | ···················· | NNNNNNNNNNNNNNNNNNNN | NNNNNNNNNNNNNNNNNNNN | 1 |
| E | 8476 hCoV-19/Spain/MD-IBV-004988/2020\|EPI_ISL_481085\|20 | ···················· | ···················· | ···················· | ···················· | NNNNNNNNNNNNNNNNNNNN | NNNNNNNNNNNNNNNNNNNN | 1 |
| E | 8477 hCoV-19/Spain/MD-IBV-004906/2020\|EPI_ISL_481086\|20 | ···················· | ···················· | ···················· | ···················· | NNNNNNNNNNNNNNNNNNNN | NNNNNNNNNNNNNNNNNNNN | 1 |
| E | 8478 hCoV-19/Spain/MD-IBV-004911/2020\|EPI_ISL_481087\|20 | ···················· | ···················· | ···················· | ···················· | NNNNNNNNNNNNNNNNNNNN | NNNNNNNNNNNNNNNNNNNN | 1 |
| E | 8479 hCoV-19/Spain/MD-IBV-004925/2020\|EPI_ISL_481088\|20 | ···················· | ···················· | ···················· | ···················· | NNNNNNNNNNNNNNNNNNNN | NNNNNNNNNNNNNNNNNNNN | 1 |
| E | 8480 hCoV-19/Spain/MD-IBV-004899/2020\|EPI_ISL_481089\|20 | ···················· | ···················· | ···················· | ···················· | NNNNNNNNNNNNNNNNNNNN | NNNNNNNNNNNNNNNNNNNN | 1 |
| E | 8481 hCoV-19/Spain/MD-IBV-004944/2020\|EPI_ISL_481090\|20 | ···················· | ···················· | ···················· | ···················· | NNNNNNNNNNNNNNNNNNNN | NNNNNNNNNNNNNNNNNNNN | 1 |
| E | 8482 hCoV-19/Spain/MD-IBV-004989/2020\|EPI_ISL_481091\|20 | ···················· | ···················· | ···················· | ···················· | NNNNNNNNNNNNNNNNNNNN | NNNNNNNNNNNNNNNNNNNN | 1 |
| E | 8484 hCoV-19/Spain/MD-IBV-004985/2020\|EPI_ISL_481093\|20 | ···················· | ···················· | ···················· | ···················· | NNNNNNNNNNNNNNNNNNNN | NNNNNNNNNNNNNNNNNNNN | 1 |
| E | 8485 hCoV-19/Spain/MD-IBV-004901/2020\|EPI_ISL_481094\|20 | ···················· | ···················· | ···················· | ···················· | NNNNNNNNNNNNNNNNNNNN | NNNNNNNNNNNNNNNNNNNN | 1 |
| E | 8486 hCoV-19/Spain/MD-IBV-004905/2020\|EPI_ISL_481096\|20 | ···················· | ···················· | ···················· | ···················· | NNNNNNNNNNNNNNNNNNNN | NNNNNNNNNNNNNNNNNNNN | 1 |
| E | 8487 hCoV-19/Spain/MD-IBV-004907/2020\|EPI_ISL_481097\|20 | ···················· | ···················· | ···················· | ···················· | NNNNNNNNNNNNNNNNNNNN | NNNNNNNNNNNNNNNNNNNN | 1 |
| E | 8488 hCoV-19/Spain/MD-IBV-004976/2020\|EPI_ISL_481098\|20 | ···················· | ···················· | ···················· | ···················· | NNNNNNNNNNNNNNNNNNNN | NNNNNNNNNNNNNNNNNNNN | 1 |
| E | 8489 hCoV-19/Spain/MD-IBV-004967/2020\|EPI_ISL_481099\|20 | ···················· | ···················· | ···················· | ···················· | NNNNNNNNNNNNNNNNNNNN | ···················· | 1 |
| E | 8505 hCoV-19/Switzerland/ZH-ETHZ-873/2020\|EPI_ISL_48368 | ···················· | ···················· | ···················· | ···················· | ······NNNNNNNNNNNNNN | NNNNNNNNNNNNNNNNNNNN | 1 |
| E | 8524 hCoV-19/USA/MI-UM-2006S001014/2020\|EPI_ISL_487230\| | ···················· | ···················· | ···················· | ···················· | NNNNNNNNNNNNNNNNNNNN | ···················· | 1 |
| E | 8590 hCoV-19/USA/FL-BPHL-0394/2020\|EPI_ISL_489803\|2020- | ···················· | ···················· | ···················· | ···················· | NNNNNNNNN··········· | ···················· | 1 |
| E | 8591 hCoV-19/USA/FL-BPHL-0401/2020\|EPI_ISL_489810\|2020- | ···················· | ···················· | ···················· | ···················· | NNNNNNNNNNNNNNNNNNNN | ···················· | 1 |
| E | 8592 hCoV-19/USA/FL-BPHL-0408/2020\|EPI_ISL_489817\|2020- | ···················· | ···················· | ···················· | ···················· | NNNNNNNNNNNN········ | ···················· | 1 |
| E | 8705 hCoV-19/Spain/PV-IBV-003202/2020\|EPI_ISL_481000\|20 | ···················· | ···················· | ···················· | ···················· | NNNNNNNNNNNNNNNNNNNN | NNNNNNNNNNNNNNNNNNNN | 1 |
| E | 8706 hCoV-19/Spain/PV-IBV-003239/2020\|EPI_ISL_481002\|20 | ···················· | ···················· | ···················· | ···················· | NNNNNNNNNNNNNNNNNNNN | NNNNNNNNNNNNNNNNNNNN | 1 |
| E | 8707 hCoV-19/Spain/CT-IBV-003734/2020\|EPI_ISL_481003\|20 | ···················· | ···················· | ···················· | ···················· | NNNNNNNNNNNNNNNNNNNN | NNNNNNNNNNNNNNNNNNNN | 1 |
| E | 8708 hCoV-19/Spain/PV-IBV-003260/2020\|EPI_ISL_481004\|20 | ···················· | ···················· | ···················· | ···················· | ·NNNNNNNNNNN········ | ···················· | 1 |
| E | 8709 hCoV-19/Spain/PV-IBV-003209/2020\|EPI_ISL_481005\|20 | ···················· | ···················· | ···················· | ···················· | NNNNNNN············· | ···················· | 1 |
| E | 8710 hCoV-19/Spain/PV-IBV-003207/2020\|EPI_ISL_481007\|20 | ···················· | ···················· | ···················· | ···················· | NNNNNNNNNNNNNNNNNNNN | NNNNNNNNNNNNNNNNNNNN | 1 |
| E | 8711 hCoV-19/Spain/PV-IBV-003243/2020\|EPI_ISL_481008\|20 | ···················· | ···················· | ···················· | ···················· | NNNNNNNNNNNNNNNNNNNN | NNNNNNNNNNNNNNNNNNNN | 1 |
| E | 8712 hCoV-19/Spain/PV-IBV-003193/2020\|EPI_ISL_481009\|20 | ···················· | ···················· | ···················· | ···················· | NNNNNNNNNNNNNNNNNNNN | NNNNNNNNNNNNNNNNNNNN | 1 |
| E | 8713 hCoV-19/Spain/PV-IBV-003198/2020\|EPI_ISL_481010\|20 | ···················· | ···················· | ···················· | ···················· | NNNNNNNNNNNNNNNNNNNN | NNNNNNNNNNNNNNNNNNNN | 1 |
| E | 8714 hCoV-19/Spain/PV-IBV-003267/2020\|EPI_ISL_481011\|20 | ···················· | ···················· | ···················· | ···················· | NNNNNNNNNNNNNNNNNNNN | NNNNNNNNNNNNNNNNNNNN | 1 |
| E | 8715 hCoV-19/Spain/PV-IBV-003189/2020\|EPI_ISL_481013\|20 | ···················· | ···················· | ···················· | ···················· | NNNNNNNNNNNNNNNNNNNN | NNNNNNNNNNNNNNNNNNNN | 1 |
| E | 8716 hCoV-19/Spain/CT-IBV-003735/2020\|EPI_ISL_481017\|20 | ···················· | ···················· | ···················· | ···················· | NNNNNNNNNNNNNNNNNNNN | NNNNNNNNNNNNNNNNNNNN | 1 |
| E | 8717 hCoV-19/Spain/PV-IBV-003199/2020\|EPI_ISL_481020\|20 | ···················· | ···················· | ···················· | ···················· | NNNNNNNNNNNNNNNNNNNN | NNNNNNNNNNNNNNNNNNNN | 1 |
| E | 8718 hCoV-19/Spain/PV-IBV-003262/2020\|EPI_ISL_481032\|20 | ···················· | ···················· | ···················· | ···················· | NNNNNNNNNNNNNNNNNNNN | NNNNNNNNNNNNNNNNNNNN | 1 |
| E | 8719 hCoV-19/Spain/PV-IBV-003216/2020\|EPI_ISL_481033\|20 | ···················· | ···················· | ···················· | ···················· | NNNNNNNNNNNNNNNNNNNN | NNNNNNNNNNNNNNNNNNNN | 1 |
| E | 8720 hCoV-19/Spain/CT-IBV-003737/2020\|EPI_ISL_481034\|20 | ···················· | ···················· | ···················· | ···················· | NNNNNNNNNNNNNNNNNNNN | NNNNNNNNNNNNNNNNNNNN | 1 |
| E | 8721 hCoV-19/Spain/PV-IBV-003206/2020\|EPI_ISL_481036\|20 | ···················· | ···················· | ···················· | ···················· | NNNNNNNNNNNNNNNNNNNN | NNNNNNNNNNNNNNNNNNNN | 1 |
| E | 8722 hCoV-19/Spain/PV-IBV-003261/2020\|EPI_ISL_481037\|20 | ···················· | ···················· | ···················· | ···················· | NNNNNNNNNNNNNNNNNNNN | NNNNNNNNNNNNNNN····· | 1 |
| E | 8723 hCoV-19/Spain/PV-IBV-003218/2020\|EPI_ISL_481038\|20 | ···················· | ···················· | ···················· | ···················· | NNNNNNNNNNNNNNNNNNNN | NNNNNNNNNNNNNNNNNNNN | 1 |
| E | 8724 hCoV-19/Spain/PV-IBV-003245/2020\|EPI_ISL_481039\|20 | ···················· | ···················· | ···················· | ···················· | NNNNNNNNNNNNNNNNNNNN | NNNNNNNNNNNNNNNNNNNN | 1 |
| E | 8725 hCoV-19/Spain/PV-IBV-003233/2020\|EPI_ISL_481040\|20 | ···················· | ···················· | ···················· | ···················· | NNNNNNNNNNNNNNNNNNNN | NNNNNNNNNNNNNNNNNNNN | 1 |
| E | 8726 hCoV-19/Spain/MD-IBV-004968/2020\|EPI_ISL_481041\|20 | ···················· | ···················· | ···················· | ···················· | NNNNNNNNNNNNNNNNNNNN | NNNNNNNNNNNNNNN····· | 1 |
| E | 8727 hCoV-19/Spain/MD-IBV-004921/2020\|EPI_ISL_481043\|20 | ···················· | ···················· | ···················· | ···················· | NNNNNNNNNNNNNNNNNNNN | N···········NNNNNNNN | 1 |
| E | 8728 hCoV-19/Spain/MD-IBV-004903/2020\|EPI_ISL_481044\|20 | ···················· | ···················· | ···················· | ···················· | NNNNNNNNNNNNNNNNNNNN | NNNNNNNNNNNNNNNNNNNN | 1 |
| E | 8729 hCoV-19/Spain/MD-IBV-004981/2020\|EPI_ISL_481045\|20 | ···················· | ···················· | ···················· | ···················· | NNNNNNNNNNNNNNNNNNNN | NNNNNNNNNNNNNNNNNNNN | 1 |
| E | 8730 hCoV-19/Spain/MD-IBV-004961/2020\|EPI_ISL_481047\|20 | ···················· | ···················· | ···················· | ···················· | NNNNNNNNNNNNNNNNNNNN | NNNNNNNNNNNNNNNNNNNN | 1 |
| E | 8731 hCoV-19/Spain/MD-IBV-004934/2020\|EPI_ISL_481048\|20 | ···················· | ···················· | ···················· | ···················· | NNNNNNNNNNNNNNNNNNNN | ···················· | 1 |
| E | 8732 hCoV-19/Spain/MD-IBV-004956/2020\|EPI_ISL_481049\|20 | ···················· | ···················· | ···················· | ···················· | NNNNNNNNNNNNNNNNNNNN | NNNNNNNNNNNNNNNNNNNN | 1 |
| E | 8733 hCoV-19/Spain/MD-IBV-004904/2020\|EPI_ISL_481051\|20 | ···················· | ···················· | ···················· | ···················· | NNNNNNNNNNNNNNNNNNNN | NNNNNNNNNNNNNNNNNNNN | 1 |
| E | 8734 hCoV-19/Spain/MD-IBV-004900/2020\|EPI_ISL_481052\|20 | ···················· | ···················· | ···················· | ···················· | NNNNNNNNNNNNNNNNNNNN | NNNNNNNNNNNNNNNNNNNN | 1 |
| E | 8735 hCoV-19/Spain/MD-IBV-004920/2020\|EPI_ISL_481054\|20 | ···················· | ···················· | ···················· | ···················· | NNNNNNNNNNNNNNNNNNNN | NNNNNNNNNNNNNNNNNNNN | 1 |
| E | 8736 hCoV-19/Spain/MD-IBV-004915/2020\|EPI_ISL_481055\|20 | ···················· | ···················· | ···················· | ···················· | NNNNNNNNNNNNNNNNNNNN | NNNNNNNNNNNNNNNNNNNN | 1 |
| E | 8737 hCoV-19/Spain/MD-IBV-004922/2020\|EPI_ISL_481056\|20 | ···················· | ···················· | ···················· | ···················· | NNNNNNNNNNNNNNNNNNNN | NNNNNNNNNNNNNNNNNNNN | 1 |
| E | 8738 hCoV-19/Spain/MD-IBV-004910/2020\|EPI_ISL_481057\|20 | ···················· | ···················· | ···················· | ···················· | NNNNNNNNNNNNNNNNNNNN | NNNNNNNNNNNNNNNNNNNN | 1 |
| E | 8739 hCoV-19/Spain/MD-IBV-004902/2020\|EPI_ISL_481058\|20 | ···················· | ···················· | ···················· | ···················· | NNNNNNNNNNNNNNNNNNNN | NNNNNNNNNNNNNNNNNNNN | 1 |
| E | 8740 hCoV-19/Spain/MD-IBV-004912/2020\|EPI_ISL_481059\|20 | ···················· | ···················· | ···················· | ···················· | ··NNNNN···NNNNNNNNNN | NNNNNNNNNNNNNNN····· | 1 |
| E | 8741 hCoV-19/Spain/MD-IBV-004917/2020\|EPI_ISL_481060\|20 | ···················· | ···················· | ···················· | ···················· | NNNNNNNNNNNNNNNNNNNN | NNNNNNNNNNNNNNNNNNNN | 1 |
| E | 8742 hCoV-19/Spain/MD-IBV-004936/2020\|EPI_ISL_481062\|20 | ···················· | ···················· | ···················· | ···················· | NNNNNNNNNNNNNNNNNNNN | NNNNN····NNNNNN····· | 1 |
| E | 8743 hCoV-19/Spain/MD-IBV-004982/2020\|EPI_ISL_481063\|20 | ···················· | ···················· | ···················· | ···················· | NNNNNNNNNNNNNNNNNNNN | NNNNNNNNNNNNNNNNNNNN | 1 |

| | | | | | | | | | |
|---|---|---|---|---|---|---|---|---|---|
| E | 8744 | hCoV-19/Spain/MD-IBV-004938/2020\|EPI_ISL_481064\|20 | ···················· | ···················· | ···················· | ···················· | NNNNNNNNNNNNNNNNNNNN | NNNNNNNNNNNNNNNNNNNN | 1 |
| E | 8837 | hCoV-19/Switzerland/ZH-ETHZ-1071/2020\|EPI_ISL_4836 | ···················· | ···················· | ···················· | ···················· | ······NNNNNNNNNNNNNN | NNNNNNNNNNNNNNNNNNNN | 1 |
| E | 8853 | hCoV-19/USA/WI-UW-607/2020\|EPI_ISL_484982\|2020-06- | ···················· | ···················· | ···················· | ···················· | NNNNNNNNNNNNNNNNNNNN | NNNNNNNNNNNNNNNNNNNN | 1 |
| E | 8854 | hCoV-19/USA/WI-UW-608/2020\|EPI_ISL_484983\|2020-06- | ···················· | ···················· | ···················· | ···················· | NNNNNNNNNNNNNNNNNNNN | NNNNNNNNNNNNNNNNNNNN | 1 |
| E | 8855 | hCoV-19/USA/WI-UW-609/2020\|EPI_ISL_484984\|2020-06- | ···················· | ···················· | ···················· | ···················· | NNNNNNNNNNNNNNNNNNNN | NNNNNNNNNNNNNNNNNNNN | 1 |
| E | 8961 | hCoV-19/Spain/PV-IBV-003232/2020\|EPI_ISL_481014\|20 | ···················· | ···················· | ···················· | ···················· | NNNNNNNNNNNNNNNNNNNN | NNNNNNNNNNNNNNNNNNNN | 1 |
| E | 8962 | hCoV-19/Spain/PV-IBV-003208/2020\|EPI_ISL_481015\|20 | ···················· | ···················· | ···················· | ···················· | NNNNNNNNNNNNNNNNNNNN | NNNNNNNNNNNNNNNNNNNN | 1 |
| E | 8963 | hCoV-19/Spain/PV-IBV-003235/2020\|EPI_ISL_481018\|20 | ···················· | ···················· | ···················· | ···················· | NNNNNNNNNNNNNNNNNNNN | NNNNNNNNNNNNNNNNNNNN | 1 |
| E | 8964 | hCoV-19/Spain/PV-IBV-003247/2020\|EPI_ISL_481019\|20 | ···················· | ···················· | ···················· | ···················· | NNNNNNN············· | ···················· | 1 |
| E | 9029 | hCoV-19/USA/WI-UW-538/2020\|EPI_ISL_484913\|2020-06- | ···················· | ···················· | ···················· | ···················· | NNNNNNNNNNNNNNNNNNNN | NNNNNNNNNNNNNNNNNNNN | 1 |
| E | 9030 | hCoV-19/USA/WI-UW-539/2020\|EPI_ISL_484914\|2020-06- | ···················· | ···················· | ···················· | ···················· | NNNNNNNNNNNNNNNNNNNN | NNNNNNNNNNNNNNNNNNNN | 1 |
| E | 9031 | hCoV-19/USA/WI-UW-540/2020\|EPI_ISL_484915\|2020-06- | ···················· | ···················· | ···················· | ···················· | NNNNNNNNNNNNNNNNNNNN | NNNNNNNNNNNNNNNNNNNN | 1 |
| E | 9034 | hCoV-19/USA/WI-UW-543/2020\|EPI_ISL_484918\|2020-06- | ···················· | ···················· | ···················· | ···················· | NNNNNNNNNNNNNNNNNNNN | NNNNNNNNNNNNNNNNNNNN | 1 |
| E | 9036 | hCoV-19/USA/WI-UW-545/2020\|EPI_ISL_484920\|2020-06- | ···················· | ···················· | ···················· | ···················· | NNNNNNNNNNNNNNNNNNNN | NNNNNNNNNNNNNNNNNNNN | 1 |
| E | 9037 | hCoV-19/USA/WI-UW-546/2020\|EPI_ISL_484921\|2020-06- | ···················· | ···················· | ···················· | ···················· | NNNNNNNNNNNNNNNNNNNN | NNNNNNNNNNNNNNNNNNNN | 1 |
| E | 9039 | hCoV-19/USA/WI-UW-548/2020\|EPI_ISL_484923\|2020-06- | ···················· | ···················· | ···················· | ···················· | NNNNNNNNNNNNNNNNNNNN | NNNNNNNNNNNNNNNNNNNN | 1 |
| E | 9040 | hCoV-19/USA/WI-UW-549/2020\|EPI_ISL_484924\|2020-06- | ···················· | ···················· | ···················· | ···················· | NNNNNNNNNNNNNNNNNNNN | NNNNNNNNNNNNNNNNNNNN | 1 |
| E | 9043 | hCoV-19/USA/IL-UW-553/2020\|EPI_ISL_484928\|2020-06- | ···················· | ·A·················· | ···················· | ···················· | NNNNNNNNNNNNNNNNNNNN | NNNNNNNNNNNNNNNNNNNN | 1 |
| E | 9046 | hCoV-19/USA/WI-UW-558/2020\|EPI_ISL_484933\|2020-06- | ···················· | ···················· | ···················· | ···················· | NNNNNNNNNNNNNNNNNNNN | NNNNNNNNNNNNNNNNNNNN | 1 |
| E | 9047 | hCoV-19/USA/WI-UW-559/2020\|EPI_ISL_484934\|2020-06- | ···················· | ···················· | ···················· | ···················· | NNNNNNNNNNNNNNNNNNNN | NNNNNNNNNNNNNNNNNNNN | 1 |
| E | 9050 | hCoV-19/USA/WI-UW-562/2020\|EPI_ISL_484937\|2020-06- | ···················· | ···················· | ···················· | ···················· | NNNNNNNNNNNNNNNNNNNN | NNNNNNNNNNNNNNNNNNNN | 1 |
| E | 9473 | hCoV-19/Spain/MD-IBV-004983/2020\|EPI_ISL_481100\|20 | ···················· | ···················· | ···················· | ···················· | NNNNNNNNNNNNNNNNNNNN | NNNNNNNNNNNNNNNNNNNN | 1 |
| E | 9474 | hCoV-19/Spain/MD-IBV-004945/2020\|EPI_ISL_481101\|20 | ···················· | ···················· | ···················· | ···················· | NNNNNNNNNNNNNNNNNNNN | NNNNNNNNNNNNNNNNNNNN | 1 |
| E | 9475 | hCoV-19/Spain/MD-IBV-004926/2020\|EPI_ISL_481102\|20 | ···················· | ···················· | ···················· | ···················· | NNNNNNNNNNNNNNNNNNNN | NNNNNNNNNNNNNNNNNNNN | 1 |
| E | 9476 | hCoV-19/Spain/MD-IBV-004919/2020\|EPI_ISL_481103\|20 | ···················· | ···················· | ···················· | ···················· | NNNNNNNNNNNNNNNNNNNN | NNNNNNNNNNNNNNNNNNNN | 1 |
| E | 9477 | hCoV-19/Spain/MD-IBV-004937/2020\|EPI_ISL_481104\|20 | ···················· | ···················· | ···················· | ···················· | NNNNNNNNNNNNNNNNNNNN | NNNNNNNNNNNNNNNNNNNN | 1 |
| E | 9478 | hCoV-19/Spain/MD-IBV-004963/2020\|EPI_ISL_481105\|20 | ···················· | ···················· | ···················· | ···················· | NNNNNNN············· | ···················· | 1 |
| E | 9479 | hCoV-19/Spain/MD-IBV-004929/2020\|EPI_ISL_481106\|20 | ···················· | ···················· | ···················· | ···················· | NNNNNNNNNNNNNNNNNNNN | NNNNNNNNNNNNNNNNNNNN | 1 |
| E | 9480 | hCoV-19/Spain/MD-IBV-004897/2020\|EPI_ISL_481107\|20 | ···················· | ···················· | ···················· | ···················· | NNNNNNNNNNNNNNNNNNNN | NNNNNNNNNNNNNNNNNNNN | 1 |
| E | 9481 | hCoV-19/Spain/MD-IBV-004980/2020\|EPI_ISL_481108\|20 | ···················· | ···················· | ···················· | ···················· | NNNNNNNNNNNNNNNNNNNN | NNNNNNNNNNNNNNNNNNNN | 1 |
| E | 9482 | hCoV-19/Spain/MD-IBV-004941/2020\|EPI_ISL_481109\|20 | ···················· | ···················· | ···················· | ···················· | NNNNNNNNNNNNNNNNNNNN | NNNNNNNNNNNNNNNNNNNN | 1 |
| E | 9670 | hCoV-19/USA/CA-ALSR-1283/2020\|EPI_ISL_494479\|2020- | ···················· | ·A·················· | ···················· | ···················· | ···········NNN···NNN | ···················· | 1 |
| E | 9676 | hCoV-19/USA/CA-ALSR-1302/2020\|EPI_ISL_494486\|2020- | ···················· | ·A·················· | ···················· | ···················· | NNNNNNNNNNNNNNNNNNNN | ···················· | 1 |
| E | 9806 | hCoV-19/South Africa/KRISP-0312/2020\|EPI_ISL_48729 | ···················· | ···················· | ···················· | ···················· | ········NNNNNNNNNN·· | ···················· | 1 |
| F | 189 | hCoV-19/USA/FL-BPHL-0724/2020\|EPI_ISL_509782\|2020- | ···················· | ···················· | ···················· | ···················· | NNNNNNNNNNNNNNNNNNNN | NNNNNNNNNNNNNNNNNNNN | 1 |
| F | 200 | hCoV-19/USA/FL-BPHL-0738/2020\|EPI_ISL_509796\|2020- | ···················· | ···················· | ···················· | ···················· | NNNNNNNNNNNNNNNNNNNN | NNNNNNNNNNNNNNNNNNNN | 1 |
| F | 323 | hCoV-19/England/LIVE-A4A3D/2020\|EPI_ISL_499812\|202 | ···················· | ···················· | ···················· | ···················· | ······NNNNNNNNNNNNNN | NNNNNNNNNNNNNNNNNNNN | 1 |
| F | 324 | hCoV-19/England/LIVE-A4AA6/2020\|EPI_ISL_499813\|202 | ···················· | ···················· | ···················· | ···················· | ······NNNNNNNNNNNNNN | NNNNNNNNNNNNNNNNNNNN | 1 |
| F | 328 | hCoV-19/England/LIVE-A4CBF/2020\|EPI_ISL_499817\|202 | ···················· | ···················· | ···················· | ···················· | ······NNNNNNNNNNNNNN | NNNNNNNNNNNNNNNNNNNN | 1 |
| F | 329 | hCoV-19/England/LIVE-A4F3E/2020\|EPI_ISL_499818\|202 | ···················· | ···················· | ···················· | ···················· | ······NNNNNNNNNNNNNN | ···················· | 1 |
| F | 331 | hCoV-19/England/LIVE-A552C/2020\|EPI_ISL_499820\|202 | ···················· | ···················· | ···················· | ···················· | ······NNNNNNNNNNNNNN | NNNNNNNNNNNNNNNNNNNN | 1 |
| F | 332 | hCoV-19/England/LIVE-A5595/2020\|EPI_ISL_499821\|202 | ···················· | ···················· | ···················· | ···················· | ······NNNNNNNNNNNNNN | ···················· | 1 |
| F | 465 | hCoV-19/Israel/CVL-n26456/2020\|EPI_ISL_514303\|2020 | ···················· | ···················· | ···················· | ···················· | ······NNNNNNNNNNNNNN | NNNNNNNNNNNNNNNNNNNN | 1 |
| F | 496 | hCoV-19/South Africa/KRISP-K002244/2020\|EPI_ISL_51 | ···················· | ···················· | ···················· | ···················· | ·NNNNNNNNNNNNNNNNNNN | ···················· | 1 |
| F | 504 | hCoV-19/South Africa/KRISP-K002314/2020\|EPI_ISL_51 | ···················· | ···················· | ···················· | ···················· | ··NNNNNNNNNNNNNNNNNN | NNNNNNNNNNNNNNNNNNNN | 1 |
| F | 587 | hCoV-19/USA/WI-UW-841/2020\|EPI_ISL_509848\|2020-05- | ···················· | ···················· | ···················· | ···················· | NNNNNNNNNNNNNNNNNNNN | NNNNNNNNNNNNNNNNNNNN | 1 |
| F | 591 | hCoV-19/USA/WI-UW-845/2020\|EPI_ISL_509852\|2020-07- | ···················· | ···················· | ···················· | ···················· | NNNNNNNNNNNNNNNNNNNN | NNNNNNNNNNNNNNNNNNNN | 1 |
| F | 597 | hCoV-19/USA/WI-UW-853/2020\|EPI_ISL_509859\|2020-07- | ···················· | ···················· | ···················· | ···················· | NNNNNNNNNNNNNNNNNNNN | NNNNNNNNNNNNNNNNNNNN | 1 |
| F | 598 | hCoV-19/USA/WI-UW-854/2020\|EPI_ISL_509860\|2020-07- | ···················· | ···················· | ···················· | ···················· | NNNNNNNNNNNNNNNNNNNN | NNNNNNNNNNNNNNNNNNNN | 1 |
| F | 600 | hCoV-19/USA/WI-UW-858/2020\|EPI_ISL_509863\|2020-07- | ···················· | ···················· | ···················· | ···················· | NNNNNNNNNNNNNNNNNNNN | NNNNNNNNNNNNNNNNNNNN | 1 |
| F | 601 | hCoV-19/USA/WI-UW-861/2020\|EPI_ISL_509864\|2020-07- | ···················· | ···················· | ···················· | ···················· | NNNNNNNNNNNNNNNNNNNN | NNNNNNNNNNNNNNNNNNNN | 1 |
| F | 602 | hCoV-19/USA/WI-UW-862/2020\|EPI_ISL_509865\|2020-07- | ···················· | ·A·················· | ···················· | ···················· | NNNNNNNNNNNNNNNNNNNN | NNNNNNNNNNNNNNNNNNNN | 1 |
| F | 603 | hCoV-19/USA/WI-UW-863/2020\|EPI_ISL_509866\|2020-07- | ···················· | ···················· | ···················· | ···················· | NNNNNNNNNNNNNNNNNNNN | NNNNNNNNNNNNNNNNNNNN | 1 |
| F | 765 | hCoV-19/South Africa/KRISP-K002309/2020\|EPI_ISL_51 | ···················· | ···················· | ···················· | ···················· | NNNNNNNNNNNNNNNNNNNN | ···················· | 1 |
| F | 766 | hCoV-19/South Africa/KRISP-K002310/2020\|EPI_ISL_51 | ··················NN | ···················· | ···················· | ···················· | ·NNNNNNNNNNNN······· | ···················· | 1 |
| F | 1161 | hCoV-19/England/LIVE-A1283/2020\|EPI_ISL_499987\|202 | ···················· | ·A·················· | ···················· | ···················· | ······NNNNNNNNNNNNNN | NNNNNNNNNNNNNNNNNNNN | 1 |
| F | 1162 | hCoV-19/England/LIVE-A1292/2020\|EPI_ISL_499988\|202 | ···················· | ·A·················· | ···················· | ···················· | ······NNNNNNNNNNNNNN | NNNNNNNNNNNNNNNNNNNN | 1 |
| F | 1163 | hCoV-19/England/LIVE-A12A1/2020\|EPI_ISL_499989\|202 | ···················· | ·A·················· | ···················· | ···················· | ······NNNNNNNNNNNNNN | NNNNNNNNNNNNNNNNNNNN | 1 |
| F | 1164 | hCoV-19/England/LIVE-A12B0/2020\|EPI_ISL_499990\|202 | ···················· | ·A·················· | ···················· | ···················· | ······NNNNNNNNNNNNNN | NNNNNNNNNNNNNNNNNNNN | 1 |
| F | 1165 | hCoV-19/England/LIVE-A12CF/2020\|EPI_ISL_499991\|202 | ···················· | ·A·················· | ···················· | ···················· | ······NNNNNNNNNNNNNN | ···················· | 1 |
| F | 1166 | hCoV-19/England/LIVE-A12ED/2020\|EPI_ISL_499992\|202 | ···················· | ·A·················· | ···················· | ···················· | ······NNNNNNNNNNNNNN | ···················· | 1 |
| F | 1167 | hCoV-19/England/LIVE-A1317/2020\|EPI_ISL_499993\|202 | ···················· | ·A·················· | ···················· | ···················· | ······NNNNNNNNNNNNNN | ···················· | 1 |
| F | 1360 | hCoV-19/England/LIVE-A120B/2020\|EPI_ISL_499984\|202 | ···················· | ·A·················· | ···················· | ···················· | ······NNNNNNNNNNNNNN | ···················· | 1 |
| F | 1370 | hCoV-19/England/LOND-124D390/2020\|EPI_ISL_507091\|2 | ···················· | ···················· | ···················· | ···················· | ·NNNNNNNNNNNNNNNNNNN | NNNNNNNNNNNNNNNNNNNN | 1 |
| F | 1373 | hCoV-19/England/LOND-124D3DC/2020\|EPI_ISL_507094\|2 | ···················· | ···················· | ···················· | ···················· | NNNNNNNNNNNNNNNNNNNN | NNNNNNNNNNNNNNNNNNNN | 1 |
| F | 1375 | hCoV-19/England/LOND-124D424/2020\|EPI_ISL_507097\|2 | ···················· | ···················· | ···················· | ···················· | ··NNNNNNNNNNNNNNNNNN | ···················· | 1 |
| F | 1482 | hCoV-19/Israel/CVL-n25870/2020\|EPI_ISL_514271\|2020 | ···················· | ···················· | ···················· | ···················· | ···NNNNNNNNNNNNNNNNN | NNNNNNNNNNNNNNNNNNNN | 1 |
| F | 1486 | hCoV-19/Israel/CVL-n25922/2020\|EPI_ISL_514278\|2020 | ···················· | ···················· | ··········T········· | ···················· | ·······NNNNNNNNNNNNN | ···················· | 1 |
| F | 1674 | hCoV-19/USA/FL-BPHL-0665/2020\|EPI_ISL_509723\|2020- | ···················· | ···················· | ···················· | ···················· | NNNNNNNNNNNNNNNNNNNN | NNNNNNNNNNNNNNNNNNNN | 1 |
| F | 1680 | hCoV-19/USA/FL-BPHL-0671/2020\|EPI_ISL_509729\|2020- | ···················· | ···················· | ···················· | ···················· | NNNNNNNNNNNNNNNNNNNN | ···················· | 1 |
| F | 1684 | hCoV-19/USA/FL-BPHL-0675/2020\|EPI_ISL_509733\|2020- | ···················· | ···················· | ···················· | ···················· | NNNNNNNNNNNNNNNNNNNN | NNNNNNNNNNNNNNNNNNNN | 1 |
| F | 1688 | hCoV-19/USA/FL-BPHL-0679/2020\|EPI_ISL_509737\|2020- | ···················· | ···················· | ···················· | ···················· | NNNNNNNNNNNNNNNNNNNN | NNNNNNNNNNNNNNNNNNNN | 1 |
| F | 1694 | hCoV-19/USA/FL-BPHL-0685/2020\|EPI_ISL_509743\|2020- | ···················· | ···················· | ···················· | ···················· | NNNNNNNNNNNNNNNNNNNN | NNNNNNNNNNNNNNNNNNNN | 1 |
| F | 1696 | hCoV-19/USA/FL-BPHL-0688/2020\|EPI_ISL_509746\|2020- | ···················· | ···················· | ···················· | ···················· | ········NNN·······NN | NNNNNNNNNNN········· | 1 |
| F | 1699 | hCoV-19/USA/FL-BPHL-0691/2020\|EPI_ISL_509749\|2020- | ···················· | ···················· | ···················· | ···················· | NNNNNNNNNNNNNNNNNNNN | NNNNNNNNNNNNNNNNNNNN | 1 |
| F | 1700 | hCoV-19/USA/FL-BPHL-0692/2020\|EPI_ISL_509750\|2020- | ···················· | ···················· | ···················· | ···················· | NNNNNNNNNNNNNN·····N | ···················· | 1 |
| F | 1785 | hCoV-19/Israel/CVL-n26140/2020\|EPI_ISL_514290\|2020 | ···················· | ···················· | ···················· | ···················· | ·······NNNNNNNNNNNNN | NNNNNNNNNNNNNNNNNNNN | 1 |
| F | 1791 | hCoV-19/Israel/CVL-n26170/2020\|EPI_ISL_514296\|2020 | ···················· | ·A·················· | ···················· | ···················· | ······NNNNNNNNNNNNNN | NNNNNNNNNNNNNNNNNNNN | 1 |
| F | 1848 | hCoV-19/England/LIVE-A5C63/2020\|EPI_ISL_499656\|202 | ···················· | ···················· | ···················· | ···················· | ······NNNNNNNNNNNNNN | NNNNNNNNNNNNNNNNNNNN | 1 |
| F | 1849 | hCoV-19/England/LIVE-A5C90/2020\|EPI_ISL_499657\|202 | ···················· | ···················· | ···················· | ···················· | ······NNNNNNNNNNNNNN | NNNNNNNNNNNNNNNNNNNN | 1 |

| | | | | | | | | |
|---|---|---|---|---|---|---|---|---|
| F | 1850 hCoV-19/England/LIVE-A5CEB/2020\|EPI_ISL_499658\|202 | ···················· | ···················· | ···················· | ···················· | ·······NNNNNNNNNNNNN | NNNNNNNNNNNNNNNNNNNN | 1 |
| F | 1854 hCoV-19/England/LIVE-A5D9D/2020\|EPI_ISL_499662\|202 | ···················· | ···················· | ···················· | ···················· | ·······NNNNNNNNNNNNN | ···················· | 1 |
| F | 1855 hCoV-19/England/LIVE-A5DE8/2020\|EPI_ISL_499663\|202 | ···················· | ···················· | ···················· | ···················· | ·······NNNNNNNNNNNNN | ···················· | 1 |
| F | 1875 hCoV-19/England/LIVE-A60DF/2020\|EPI_ISL_499683\|202 | ···················· | ···················· | ···················· | ···················· | ·······NNNNNNNNNNNNN | NNNNNNNNNNNNNNNNNNNN | 1 |
| F | 1880 hCoV-19/England/LIVE-A6172/2020\|EPI_ISL_499688\|202 | ···················· | ···················· | ···················· | ···················· | ·······NNNNNNNNNNNNN | ···················· | 1 |
| F | 1948 hCoV-19/Spain/CT-IBV-005487/2020\|EPI_ISL_510523\|20 | ···················· | ·A·················· | ···················· | ···················· | NNNNNNNNNNNNNNNNNNNN | NNNNNNNNNNNNNNNNNNNN | 1 |
| F | 2049 hCoV-19/England/LIVE-A58D8/2020\|EPI_ISL_499644\|202 | ···················· | ·A·················· | ···················· | ···················· | ·······NNNNNNNNNNNNN | NNNNNNNNNNNNNNNNNNNN | 1 |
| F | 2052 hCoV-19/England/LIVE-A5911/2020\|EPI_ISL_499647\|202 | ···················· | ···················· | ···················· | ···················· | ·······NNNNNNNNNNNNN | NNNNNNNNNNNNNNNNNNNN | 1 |
| F | 2105 hCoV-19/Spain/PV-IBV-005156/2020\|EPI_ISL_509643\|20 | ···················· | ···················· | ···················· | ···················· | NNNNNNNNNNNNNNNNNNNN | ···················· | 1 |
| F | 2189 hCoV-19/USA/FL-BPHL-0795/2020\|EPI_ISL_514141\|2020- | ···················· | ···················· | ···················· | ···················· | NNNNNNNNNNNNNNNNNNNN | NNNNNNNNNNNNNNNNNNNN | 1 |
| F | 2190 hCoV-19/USA/FL-BPHL-0796/2020\|EPI_ISL_514142\|2020- | ···················· | ···················· | ···················· | ···················· | NNNNNNNNNNNNNNNNNNNN | ···················· | 1 |
| F | 2192 hCoV-19/USA/FL-BPHL-0798/2020\|EPI_ISL_514144\|2020- | ···················· | ···················· | ···················· | ···················· | NNNNNNNNNNNNNNNNNNNN | ···················· | 1 |
| F | 2195 hCoV-19/USA/FL-BPHL-0801/2020\|EPI_ISL_514147\|2020- | ···················· | ···················· | ···················· | ···················· | NNNNNNNNNNNNNNNNNNNN | NNNNNNNNNNNNNNNNNNNN | 1 |
| F | 2196 hCoV-19/USA/FL-BPHL-0802/2020\|EPI_ISL_514148\|2020- | ···················· | ···················· | ···················· | ···················· | NNNNNNNNNNNNNNNNNNNN | NNNNNNNNNNNNNNNNNNNN | 1 |
| F | 2197 hCoV-19/USA/FL-BPHL-0805/2020\|EPI_ISL_514151\|2020- | ···················· | ···················· | ···················· | ···················· | NNNNNNNNNNNNNNNNNNNN | NNNNNNNNNNNNNNNNNNNN | 1 |
| F | 2203 hCoV-19/USA/FL-BPHL-0812/2020\|EPI_ISL_514158\|2020- | ···················· | ···················· | ···················· | ···················· | NNNNNNNNNNNN········ | ···················· | 1 |
| F | 2282 hCoV-19/USA/SC-20-1167-T07/2020\|EPI_ISL_516752\|202 | ···················· | ···················· | ···················· | ······NNNNNNNNNNNN·· | ···················· | ···················· | 1 |
| F | 2294 hCoV-19/USA/SC-20-1568-T07/2020\|EPI_ISL_516768\|202 | ···················· | ···················· | ···················· | ······NNNNNNNNNNNN·· | ···················· | ···················· | 1 |
| F | 2295 hCoV-19/USA/SC-20-1577-T07/2020\|EPI_ISL_516770\|202 | ···················· | ···················· | ···················· | ··············NNNN·· | ···················· | ···················· | 1 |
| F | 2304 hCoV-19/USA/SC-20-1936-T00/2020\|EPI_ISL_516779\|202 | ···················· | ···················· | ···················· | ······NNNNNNNNNNNN·· | ···················· | ···················· | 1 |
| F | 2376 hCoV-19/Australia/SAP453/2020\|EPI_ISL_508143\|2020- | ···················· | ···················· | ···················· | ···················· | NNNNNNNNNNNNNNNNNNNN | ···················· | 1 |
| F | 2459 hCoV-19/Spain/AN-IBV-006091/2020\|EPI_ISL_510427\|20 | ···················· | ···················· | ···················· | ···················· | NNNNNNNNNNNNNNNNNNNN | NNNNNNNNNNNNNNNNNNNN | 1 |
| F | 2460 hCoV-19/Spain/AN-IBV-006095/2020\|EPI_ISL_510428\|20 | ···················· | ···················· | ···················· | ···················· | NNNNNNNNNNNNNNNNNNNN | NNNNNNNNNNNNNNNNNNNN | 1 |
| F | 2461 hCoV-19/Spain/AN-IBV-006103/2020\|EPI_ISL_510435\|20 | ···················· | ···················· | ···················· | ···················· | NNNNNNNNNNNNNNNNNNNN | NNNNNNNNNNNNNNNNNNNN | 1 |
| F | 2462 hCoV-19/Spain/AN-IBV-006104/2020\|EPI_ISL_510436\|20 | ···················· | ···················· | ···················· | ···················· | NNNNNNNNNNNNNNNNNNNN | NNNNNNNNNNNNNNNNNNNN | 1 |
| F | 2463 hCoV-19/Spain/AN-IBV-006105/2020\|EPI_ISL_510437\|20 | ···················· | ···················· | ···················· | ···················· | NNNNNNNNNNNNNNNNNNNN | NNNNNNNNNNNNNNNNNNNN | 1 |
| F | 2464 hCoV-19/Spain/AN-IBV-006108/2020\|EPI_ISL_510439\|20 | ···················· | ···················· | ···················· | ···················· | ··NNNNN············· | ···················· | 1 |
| F | 2465 hCoV-19/Spain/AN-IBV-006110/2020\|EPI_ISL_510440\|20 | ···················· | ···················· | ···················· | ···················· | NNNNNNNNNNNNNNNNNNNN | NNNNNNNNNNNNNNN····· | 1 |
| F | 2466 hCoV-19/Spain/AN-IBV-006112/2020\|EPI_ISL_510441\|20 | ···················· | ···················· | ···················· | ···················· | NNNNNNNNNNNNNNNNNNNN | NNNNNNNNNNNNNNNNNNNN | 1 |
| F | 2467 hCoV-19/Spain/AN-IBV-006115/2020\|EPI_ISL_510444\|20 | ···················· | ···················· | ···················· | ···················· | NNNNNNNNNNNNNNNNNNNN | NNNNNNNNNNNNNNN····· | 1 |
| F | 2469 hCoV-19/Spain/AN-IBV-006117/2020\|EPI_ISL_510446\|20 | ···················· | ···················· | ···················· | ···················· | NNNNNNNNNNNNNNNNNNNN | NNNNNNNNNNNNNNNNNNNN | 1 |
| F | 2470 hCoV-19/Spain/AN-IBV-006118/2020\|EPI_ISL_510447\|20 | ···················· | ···················· | ···················· | ···················· | ··NNNNN············· | ···················· | 1 |
| F | 2471 hCoV-19/Spain/AN-IBV-006119/2020\|EPI_ISL_510448\|20 | ···················· | ···················· | ···················· | ···················· | NNNNNNNNNNNN········ | ···················· | 1 |
| F | 2472 hCoV-19/Spain/AN-IBV-006120/2020\|EPI_ISL_510449\|20 | ···················· | ···················· | ···················· | ···················· | NNNNNNNNNNNNNNNNNNNN | NNNNNNNNNNNNNNNNNNNN | 1 |
| F | 2473 hCoV-19/Spain/AN-IBV-006121/2020\|EPI_ISL_510450\|20 | ···················· | ···················· | ···················· | ···················· | NNNNNNNNNNNNNNNNNNNN | NNNNNNNNNNNNNNNNNNNN | 1 |
| F | 2474 hCoV-19/Spain/AN-IBV-006122/2020\|EPI_ISL_510451\|20 | ···················· | ···················· | ···················· | ···················· | ··NNNNN············· | ····N··············· | 1 |
| F | 2476 hCoV-19/Spain/AN-IBV-006126/2020\|EPI_ISL_510454\|20 | ···················· | ···················· | ···················· | ···················· | ··NNNNN············· | ···················· | 1 |
| F | 2477 hCoV-19/Spain/AN-IBV-006128/2020\|EPI_ISL_510455\|20 | ···················· | ···················· | ···················· | ···················· | NNNNNNNNNNNNNNNNNNNN | NNNNNNNNNNNNNNNNNNNN | 1 |
| F | 2479 hCoV-19/Spain/AN-IBV-006133/2020\|EPI_ISL_510458\|20 | ···················· | ···················· | ···················· | ···················· | NNNNNNNNNNNNNNNNNNNN | NNNNNNNNNNNNNNNNNNNN | 1 |
| F | 2480 hCoV-19/Spain/AN-IBV-006134/2020\|EPI_ISL_510459\|20 | ···················· | ···················· | ···················· | ···················· | NNNNNNNNNNNNNNNNNNNN | NNNNNNNNNNNNNNNNNNNN | 1 |
| F | 2481 hCoV-19/Spain/AN-IBV-006136/2020\|EPI_ISL_510460\|20 | ···················· | ···················· | ···················· | ···················· | NNNNNNNNNNNNNNNNNNNN | NNNNNNNNNNNNNNNNNNNN | 1 |
| F | 2482 hCoV-19/Spain/CT-IBV-004174/2020\|EPI_ISL_510463\|20 | ···················· | ···················· | ···················· | ···················· | NNNNNNNNNNNNNNNNNNNN | NNNNNNNNNNNNNNNNNNNN | 1 |
| F | 2483 hCoV-19/Spain/CT-IBV-004175/2020\|EPI_ISL_510464\|20 | ···················· | ···················· | ···················· | ···················· | NNNNNNNNNNNNNNNNNNNN | NNNNNNNNNNNNNNNNNNNN | 1 |
| F | 2484 hCoV-19/Spain/CT-IBV-004176/2020\|EPI_ISL_510465\|20 | ···················· | ···················· | ···················· | ···················· | NNNNNNNNNNNNNNNNNNNN | ···················· | 1 |
| F | 2487 hCoV-19/Spain/PV-IBV-004182/2020\|EPI_ISL_510469\|20 | ···················· | ···················· | ···················· | ···················· | NNNNNNN············· | ···················· | 1 |
| F | 2490 hCoV-19/Spain/PV-IBV-004186/2020\|EPI_ISL_510473\|20 | ···················· | ···················· | ···················· | ···················· | ··NNNNNNNNNN·NNNNNNN | ···················· | 1 |
| F | 2492 hCoV-19/Spain/PV-IBV-004188/2020\|EPI_ISL_510475\|20 | ···················· | ···················· | ···················· | ···················· | NNNNNNNNNNNNNNNNNNNN | ···················· | 1 |
| F | 2493 hCoV-19/Spain/PV-IBV-004189/2020\|EPI_ISL_510476\|20 | ···················· | ···················· | ···················· | ···················· | NNNNNNNNNNNNNNNNNNNN | NNNNNNNNNNNNNNNNNNNN | 1 |
| F | 2494 hCoV-19/Spain/PV-IBV-004190/2020\|EPI_ISL_510477\|20 | ···················· | ···················· | ···················· | ···················· | NNNNNNNNNNNNNNNNNNNN | NNNNNNNNNNNNNNNNNNNN | 1 |
| F | 2495 hCoV-19/Spain/PV-IBV-004191/2020\|EPI_ISL_510478\|20 | ···················· | ···················· | ···················· | ···················· | NNNNNNNNNNNNNNNNNNNN | NNNNNNNNNNNNNNNNNNNN | 1 |
| F | 2496 hCoV-19/Spain/PV-IBV-004193/2020\|EPI_ISL_510479\|20 | ···················· | ···················· | ···················· | ···················· | NNNNNNNNNNNNNNNNNNNN | NNNNNNNNNNNNNNN·NNNN | 1 |
| F | 2497 hCoV-19/Spain/PV-IBV-004194/2020\|EPI_ISL_510480\|20 | ···················· | ···················· | ···················· | ···················· | NNNNNNNNNNNNNNNNNNNN | NNNNNNNNNNNNNNNNNNNN | 1 |
| F | 2499 hCoV-19/Spain/PV-IBV-004197/2020\|EPI_ISL_510482\|20 | ···················· | ···················· | ···················· | ···················· | NNNNNNNNNNNNNNNNNNNN | NNNNNNNNNNNNNNNNNNNN | 1 |
| F | 2500 hCoV-19/Spain/PV-IBV-004198/2020\|EPI_ISL_510483\|20 | ···················· | ···················· | ···················· | ···················· | NNNNNNNNNNNNNNNNNNNN | NNNNNNNNNNNNNNNNNNNN | 1 |
| F | 2501 hCoV-19/Spain/PV-IBV-004199/2020\|EPI_ISL_510484\|20 | ···················· | ···················· | ···················· | ···················· | NNNNNNNNNNNNNNNNNNNN | NNNNNNNNNNNNNNNNNNNN | 1 |
| F | 2502 hCoV-19/Spain/PV-IBV-004200/2020\|EPI_ISL_510485\|20 | ···················· | ···················· | ···················· | ···················· | NNNNNNNNNNNNNNNNNNNN | ···················· | 1 |
| F | 2503 hCoV-19/Spain/PV-IBV-004201/2020\|EPI_ISL_510486\|20 | ···················· | ···················· | ···················· | ···················· | NNNNNNNNNNNNNNNNNNNN | NNNNNNNNNNNNNNNNNNNN | 1 |
| F | 2504 hCoV-19/Spain/PV-IBV-004202/2020\|EPI_ISL_510487\|20 | ···················· | ···················· | ···················· | ···················· | NNNNNNNNNNNNNNNNNNNN | NNNNNNNNNNNNNNNNNNNN | 1 |
| F | 2505 hCoV-19/Spain/PV-IBV-004203/2020\|EPI_ISL_510488\|20 | ···················· | ···················· | ···················· | ···················· | NNNNNNNNNNNNNNNNNNNN | NNNNNNNNNNNNNNNNNNNN | 1 |
| F | 2506 hCoV-19/Spain/PV-IBV-004204/2020\|EPI_ISL_510489\|20 | ···················· | ···················· | ···················· | ···················· | NNNNNNNNNNNN·NNNNNNN | ···················· | 1 |
| F | 2651 hCoV-19/England/LIVE-A660A/2020\|EPI_ISL_499725\|202 | ···················· | ···················· | ···················· | ···················· | ·······NNNNNNNNNNNNN | NNNNNNNNNNNNNNNNNNNN | 1 |
| F | 2679 hCoV-19/England/LIVE-A6910/2020\|EPI_ISL_499756\|202 | ···················· | ···················· | ···················· | ···················· | ·······NNNNNNNNNNNNN | ···················· | 1 |
| F | 2697 hCoV-19/England/LIVE-A6B29/2020\|EPI_ISL_499774\|202 | ···················· | ···················· | ···················· | ···················· | ·······NNNNNNNNNNNNN | ···················· | 1 |
| F | 2768 hCoV-19/Spain/PV-IBV-004286/2020\|EPI_ISL_510402\|20 | ···················· | ···················· | ···················· | ···················· | NNNNNNNNNNNNNNNNNNNN | NNNNNNNNNNNNNNNNNNNN | 1 |
| F | 2769 hCoV-19/Spain/PV-IBV-004287/2020\|EPI_ISL_510403\|20 | ···················· | ···················· | ···················· | ···················· | NNNNNNNNNNNNNNNNNNNN | ···················· | 1 |
| F | 2774 hCoV-19/Spain/PV-IBV-004293/2020\|EPI_ISL_510409\|20 | ···················· | ···················· | ···················· | ···················· | NNNNNNNNNNNNNNNNNNNN | NNNNNNNNNNNNNNNNNNNN | 1 |
| F | 2775 hCoV-19/Spain/PV-IBV-004295/2020\|EPI_ISL_510410\|20 | ···················· | ···················· | ···················· | ···················· | NNNNNNNNNNNNNNNNNNNN | NNNNNNNNNNNNNNNNNNNN | 1 |
| F | 2776 hCoV-19/Spain/PV-IBV-004296/2020\|EPI_ISL_510411\|20 | ···················· | ···················· | ···················· | ···················· | NNNNNNNNNNNNNNNNNNNN | NNNNNNNNNNNNNNNNNNNN | 1 |
| F | 2777 hCoV-19/Spain/PV-IBV-004298/2020\|EPI_ISL_510413\|20 | ···················· | ···················· | ···················· | ···················· | NNNNNNNNNNNNNNNNN··· | ···················· | 1 |
| F | 2778 hCoV-19/Spain/PV-IBV-004300/2020\|EPI_ISL_510415\|20 | ···················· | ···················· | ···················· | ···················· | NNNNNNNNNNNNNNNNNNNN | NNNNNNNNNNNNNNNNNNNN | 1 |
| F | 2779 hCoV-19/Spain/PV-IBV-004301/2020\|EPI_ISL_510416\|20 | ···················· | ···················· | ···················· | ···················· | NNNNNNNNNNNNNNNNNNNN | NNNNNNNNNNNNNNNNNNNN | 1 |
| F | 2780 hCoV-19/Spain/PV-IBV-004303/2020\|EPI_ISL_510418\|20 | ···················· | ···················· | ···················· | ···················· | NNNNNNNNNNNNNNNNNNNN | NNNNNNNNNNNNNNNNNNNN | 1 |
| F | 2781 hCoV-19/Spain/PV-IBV-004304/2020\|EPI_ISL_510419\|20 | ···················· | ···················· | ···················· | ···················· | NNNNNNNNNNNNNNNNNNNN | ···················· | 1 |
| F | 2782 hCoV-19/Spain/PV-IBV-004305/2020\|EPI_ISL_510420\|20 | ···················· | ···················· | ···················· | ···················· | NNNNNNNNNNNNNNNNNNNN | NNNNNNNNNNNNNNN··NNN | 1 |
| F | 2783 hCoV-19/Spain/PV-IBV-004306/2020\|EPI_ISL_510421\|20 | ···················· | ···················· | ···················· | ···················· | ··NNNNN············· | ···················· | 1 |
| F | 2784 hCoV-19/Spain/PV-IBV-004307/2020\|EPI_ISL_510422\|20 | ···················· | ···················· | ···················· | ···················· | NNNNNNNNNNNNNNNNNNNN | ·············NN····· | 1 |
| F | 2785 hCoV-19/Spain/PV-IBV-004308/2020\|EPI_ISL_510423\|20 | ···················· | ···················· | ···················· | ···················· | NNNNNNNNNNNNNNNNNNNN | NNNNNNNNNNNNNNNNNNNN | 1 |
| F | 2786 hCoV-19/Spain/AN-IBV-006098/2020\|EPI_ISL_510431\|20 | ···················· | ···················· | ···················· | ···················· | NNNNNNN············· | NNNNNNN············· | 1 |
| F | 2787 hCoV-19/Spain/AN-IBV-006101/2020\|EPI_ISL_510433\|20 | ···················· | ···················· | ···················· | ···················· | NNNNNNNNNNNN········ | ···················· | 1 |
| F | 2788 hCoV-19/Spain/AN-IBV-006102/2020\|EPI_ISL_510434\|20 | ···················· | ···················· | ···················· | ···················· | NNNNNNN············· | ···················· | 1 |

| | | | | | | | | |
|---|---|---|---|---|---|---|---|---|
| F | 2870 hCoV-19/Spain/PV-IBV-004205/2020\|EPI_ISL_510490\|20 | ···················· | ···················· | ···················· | ···················· | NNNNNNNNNNNNNNNNNNNN | NNNNNNNNNNNNNNNNNNNN | 1 |
| F | 2871 hCoV-19/Spain/PV-IBV-004206/2020\|EPI_ISL_510491\|20 | ···················· | ···················· | ···················· | ···················· | NNNNNNNNNNNNNNNNNNNN | NNNNNNNNNNNNNNNNNNNN | 1 |
| F | 2872 hCoV-19/Spain/PV-IBV-004207/2020\|EPI_ISL_510492\|20 | ···················· | ···················· | ···················· | ···················· | NNNNNNNNNNNNNNNNNNNN | NNNNNNNNNNNNNNNNNNNN | 1 |
| F | 2873 hCoV-19/Spain/PV-IBV-004208/2020\|EPI_ISL_510493\|20 | ···················· | ···················· | ···················· | ···················· | NNNNNNNNNNNNNNNNNNNN | NNN················N | 1 |
| F | 2874 hCoV-19/Spain/PV-IBV-004210/2020\|EPI_ISL_510495\|20 | ···················· | ···················· | ···················· | ···················· | NNNNNNNNNNNNNNNNNNNN | ···················· | 1 |
| F | 2875 hCoV-19/Spain/PV-IBV-004211/2020\|EPI_ISL_510496\|20 | ···················· | ···················· | ···················· | ···················· | NNNNNNNNNNNNNNNNNNNN | NNNNNNNNNNNNNNNNNNNN | 1 |
| F | 2876 hCoV-19/Spain/PV-IBV-004212/2020\|EPI_ISL_510497\|20 | ···················· | ···················· | ···················· | ···················· | NNNNNNNNNNNNNNNNNNNN | NNNNNNNNNNNNNNNN···· | 1 |
| F | 2877 hCoV-19/Spain/PV-IBV-004213/2020\|EPI_ISL_510498\|20 | ···················· | ···················· | ···················· | ···················· | NNNNNNNNNNNNNNNNNNNN | ···················· | 1 |
| F | 2878 hCoV-19/Spain/PV-IBV-004214/2020\|EPI_ISL_510499\|20 | ···················· | ···················· | ···················· | ···················· | NNNNNNNNNNNNNNNNNNNN | NNNNNNNNNNNNNNNNNNNN | 1 |
| F | 2880 hCoV-19/Spain/PV-IBV-004216/2020\|EPI_ISL_510501\|20 | ···················· | ···················· | ···················· | ···················· | NNNNNNNNNNNNNNNNNNNN | ···················· | 1 |
| F | 2881 hCoV-19/Spain/PV-IBV-004217/2020\|EPI_ISL_510502\|20 | ···················· | ···················· | ···················· | ···················· | NNNNNNNNNNNNNNNNNNNN | NNNNNNNNNNNNNNNNNNNN | 1 |
| F | 2883 hCoV-19/Spain/PV-IBV-004220/2020\|EPI_ISL_510504\|20 | ···················· | ···················· | ···················· | ···················· | NNNNNNNNNNNNNNNNNNNN | ···················· | 1 |
| F | 2884 hCoV-19/Spain/PV-IBV-004222/2020\|EPI_ISL_510505\|20 | ···················· | ···················· | ···················· | ···················· | ·NNNNNN····NNNNNN··· | ·············NN····· | 1 |
| F | 2885 hCoV-19/Spain/PV-IBV-004223/2020\|EPI_ISL_510506\|20 | ···················· | ···················· | ···················· | ···················· | NNNNNNNNNNNNNNNNNNNN | NNNNNNNNNNNNNNNNNNNN | 1 |
| F | 2887 hCoV-19/Spain/PV-IBV-004226/2020\|EPI_ISL_510508\|20 | ···················· | ···················· | ···················· | ···················· | NNNNNNN············· | ···················· | 1 |
| F | 2889 hCoV-19/Spain/CT-IBV-005475/2020\|EPI_ISL_510510\|20 | ···················· | ···················· | ···················· | ···················· | NNNNNNNNNNNNNNNNNNNN | NNNNNNNNNNNNNNNNNNNN | 1 |
| F | 2890 hCoV-19/Spain/CT-IBV-005476/2020\|EPI_ISL_510511\|20 | ···················· | ···················· | ···················· | ···················· | NNNNNNNNNNNNNNNNNNNN | NNNNNNNNNNNNNNNNNNNN | 1 |
| F | 2891 hCoV-19/Spain/CT-IBV-005477/2020\|EPI_ISL_510512\|20 | ···················· | ···················· | ···················· | ···················· | NNNNNNNNNNNNNNNNNNNN | NNNNNNNNNN·········· | 1 |
| F | 2892 hCoV-19/Spain/CT-IBV-005478/2020\|EPI_ISL_510513\|20 | ···················· | ···················· | ···················· | ···················· | NNNNNNN············· | ···················· | 1 |
| F | 2893 hCoV-19/Spain/CT-IBV-005483/2020\|EPI_ISL_510514\|20 | ···················· | ·R·················· | ···················· | ···················· | NNNNNNNNNNNNNNNNNNNN | ···················· | 1 |
| F | 2894 hCoV-19/Spain/CT-IBV-005484/2020\|EPI_ISL_510515\|20 | ···················· | ···················· | ···················· | ···················· | NNNNNNNNNNNNNNNNNNNN | ···················· | 1 |
| F | 2895 hCoV-19/Spain/CT-IBV-005488/2020\|EPI_ISL_510516\|20 | ···················· | ···················· | ···················· | ···················· | ·NNNNNNNNNNNNNNNNNNN | NNNNNNNNNNNNNNNNNNNN | 1 |
| F | 2897 hCoV-19/Spain/CT-IBV-005491/2020\|EPI_ISL_510518\|20 | ···················· | ·A·················· | ···················· | ···················· | NNNNNNNNNNNNNNNNNNNN | NNNNNNNNNNNNNNNNNNNN | 1 |
| F | 2899 hCoV-19/Spain/CT-IBV-005479/2020\|EPI_ISL_510522\|20 | ···················· | ···················· | ···················· | ···················· | NNNNNNNNNNNN········ | ···················· | 1 |
| F | 3049 hCoV-19/USA/VA-DCLS-0886/2020\|EPI_ISL_516675\|2020- | ···················· | ···················A | ···················· | ···················· | ··NNNNNNNNNNNNNNNNNN | ···················· | 1 |
| F | 3133 hCoV-19/England/LIVE-A51FB/2020\|EPI_ISL_500111\|202 | ···················· | ···················· | ···················· | ···················· | ···················· | NNNNNNNNNNNNNNNNNNNN | 1 |
| F | 3136 hCoV-19/England/LIVE-A53C8/2020\|EPI_ISL_500114\|202 | ···················· | ···················· | ···················· | ···················· | ·······NNNNNNNNNNNNN | NNNNNNNNNNNNNNNNNNNN | 1 |
| F | 3139 hCoV-19/England/LIVE-A55A4/2020\|EPI_ISL_500128\|202 | ···················· | ···················· | ···················· | ···················· | ·······NNNNNNNNNNNNN | ···················· | 1 |
| F | 3140 hCoV-19/England/LIVE-A55B3/2020\|EPI_ISL_500129\|202 | ···················· | ···················· | ···················· | ···················· | ·······NNNNNNNNNNNNN | NNNNNNNNNNNNNNNNNNNN | 1 |
| F | 3148 hCoV-19/England/LIVE-A5683/2020\|EPI_ISL_500137\|202 | ···················· | ···················· | ···················· | ···················· | ·······NNNNNNNNNNNNN | NNNNNNNNNNNNNNNNNNNN | 1 |
| F | 3149 hCoV-19/England/LIVE-A5692/2020\|EPI_ISL_500138\|202 | ···················· | ···················· | ···················· | ···················· | ·······NNNNNNNNNNNNN | NNNNNNNNNNNNNNNNNNNN | 1 |
| F | 3152 hCoV-19/England/LIVE-A56DE/2020\|EPI_ISL_500141\|202 | ···················· | ···················· | ···················· | ···················· | ·······NNNNNNNNNNNNN | NNNNNNNNNNNNNNNNNNNN | 1 |
| F | 3153 hCoV-19/England/LIVE-A56ED/2020\|EPI_ISL_500142\|202 | ···················· | ···················· | ···················· | ···················· | ·······NNNNNNNNNNNNN | NNNNNNNNNNNNNNNNNNNN | 1 |
| F | 3154 hCoV-19/England/LIVE-A5708/2020\|EPI_ISL_500143\|202 | ···················· | ···················· | ···················· | ···················· | ·······NNNNNNNNNNNNN | NNNNNNNNNNNNNNNNNNNN | 1 |
| F | 3155 hCoV-19/England/LIVE-A5717/2020\|EPI_ISL_500144\|202 | ···················· | ···················· | ···················· | ···················· | ·······NNNNNNNNNNNNN | ···················· | 1 |
| F | 3156 hCoV-19/England/LIVE-A5726/2020\|EPI_ISL_500145\|202 | ···················· | ···················· | ···················· | ···················· | ·······NNNNNNNNNNNNN | ···················· | 1 |
| F | 3157 hCoV-19/England/LIVE-A5735/2020\|EPI_ISL_500146\|202 | ···················· | ···················· | ···················· | ···················· | ·············NNNNNNN | ···················· | 1 |
| F | 3160 hCoV-19/England/LIVE-A579F/2020\|EPI_ISL_500149\|202 | ···················· | ···················· | ···················· | ···················· | ·······NNNNNNNNNNNNN | NNNNNNNNNNNNNNNNNNNN | 1 |
| F | 3162 hCoV-19/England/LIVE-A57BD/2020\|EPI_ISL_500151\|202 | ···················· | ···················· | ···················· | ···················· | ·······NNNNNNNNNNNNN | NNNNNNNNNNNNN······· | 1 |
| F | 3165 hCoV-19/England/LIVE-A57F9/2020\|EPI_ISL_500154\|202 | ···················· | ···················· | ···················· | ···················· | ·······NNNNNNNNNNNNN | ···················· | 1 |
| F | 3167 hCoV-19/England/LIVE-A5832/2020\|EPI_ISL_500156\|202 | ···················· | ···················· | ···················· | ···················· | ·······NNNNNNNNNNNNN | ···················· | 1 |
| F | 3237 hCoV-19/USA/ME-HETL-J0066/2020\|EPI_ISL_513477\|2020 | ···················· | ···················· | ···················· | ···················· | ···················· | NNNNNNNN············ | 1 |
| F | 3421 hCoV-19/England/LIVE-A50B2/2020\|EPI_ISL_500104\|202 | ···················· | ···················· | ···················· | ···················· | ·······NNNNNNNNNNNNN | NNNNNNNNNNNNNNNNNNNN | 1 |
| F | 3423 hCoV-19/England/LIVE-A546B/2020\|EPI_ISL_500117\|202 | ···················· | ···················· | ···················· | ···················· | ·······NNNNNNNNNNNNN | NNNNNNNNNNNNNNNNNNNN | 1 |
| F | 3424 hCoV-19/England/LIVE-A5489/2020\|EPI_ISL_500118\|202 | ···················· | ···················· | ···················· | ···················· | ·······NNNNNNNNNNNNN | NNNNNNNNNNNNNNNNNNNN | 1 |
| F | 3426 hCoV-19/England/LIVE-A54A7/2020\|EPI_ISL_500120\|202 | ···················· | ···················· | ···················· | ···················· | ·······NNNNNNNNNNNNN | NNNNNNNNNNNNNNNNNNNN | 1 |
| F | 3429 hCoV-19/England/LIVE-A54F2/2020\|EPI_ISL_500123\|202 | ···················· | ···················· | ···················· | ···················· | ·······NNNNNNNNNNNNN | NNNNNNNNNNNNNNNNNNNN | 1 |
| F | 3430 hCoV-19/England/LIVE-A554A/2020\|EPI_ISL_500124\|202 | ···················· | ···················· | ···················· | ···················· | ·······NNNNNNNNNNNNN | ···················· | 1 |
| F | 3432 hCoV-19/England/LIVE-A5568/2020\|EPI_ISL_500126\|202 | ···················· | ···················· | ···················· | ···················· | ·······NNNNNNNNNNNNN | NNNNNNNNNNNNNNNNNNNN | 1 |
| F | 3598 hCoV-19/England/LIVE-A28F9/2020\|EPI_ISL_500011\|202 | ···················· | ···················· | ···················· | ···················· | ·······NNNNNNNNNNNNN | NNNNNNNNNNNNNNNNNNNN | 1 |
| F | 3601 hCoV-19/England/LIVE-A299C/2020\|EPI_ISL_500014\|202 | ···················· | ···················· | ···················· | ···················· | ·······NNNNNNNNNNNNN | NNNNNNNNNNNNNNNNNNNN | 1 |
| F | 3602 hCoV-19/England/LIVE-A2A7B/2020\|EPI_ISL_500015\|202 | ···················· | ···················· | ···················· | ···················· | ·······NNNNNNNNNNNN· | ···················· | 1 |
| F | 3605 hCoV-19/England/LIVE-A3C65/2020\|EPI_ISL_500018\|202 | ···················· | ···················· | ···················· | ···················· | ·······NNNNNNNNNNNNN | NNNNNNNNNNNNNNNNNNNN | 1 |
| F | 3612 hCoV-19/England/LIVE-A3D53/2020\|EPI_ISL_500025\|202 | ···················· | ···················· | ···················· | ···················· | ·······NNNNNNNNNNNNN | ···················· | 1 |
| F | 3617 hCoV-19/England/LIVE-A44C6/2020\|EPI_ISL_500030\|202 | ···················· | ···················· | ···················· | ···················· | ·······NNNNNNNNNNNNN | NNNNNNNNNNNNNNNNNNNN | 1 |
| F | 3619 hCoV-19/England/LIVE-A461B/2020\|EPI_ISL_500032\|202 | ···················· | ···················· | ···················· | ···················· | ·······NNNNNNNNNNNNN | ···················· | 1 |
| F | 3620 hCoV-19/England/LIVE-A46EE/2020\|EPI_ISL_500033\|202 | ···················· | ···················· | ···················· | ···················· | ·······NNNNNNNNNNNNN | ···················· | 1 |
| F | 3621 hCoV-19/England/LIVE-A4709/2020\|EPI_ISL_500034\|202 | ···················· | ···················· | ···················· | ···················· | ···················· | NNNN················ | 1 |
| F | 3622 hCoV-19/England/LIVE-A4718/2020\|EPI_ISL_500035\|202 | ···················· | ···················· | ···················· | ···················· | ·······NNNNNNNNNNNNN | ···················· | 1 |
| F | 3623 hCoV-19/England/LIVE-A4727/2020\|EPI_ISL_500036\|202 | ···················· | ···················· | ···················· | ···················· | ·······NNNNNNNNNNNNN | NNNNNNNNNNNNNNNNNNNN | 1 |
| F | 3626 hCoV-19/England/LIVE-A4754/2020\|EPI_ISL_500039\|202 | ···················· | ·············A······ | ···················· | ···················· | ·······NNNNNNNNNNNNN | ···················· | 1 |
| F | 3628 hCoV-19/England/LIVE-A4790/2020\|EPI_ISL_500041\|202 | ···················· | ···················· | ···················· | ···················· | ·······NNNNNNNNNNNNN | NNNNNNNNNNNNNNNNNNNN | 1 |
| F | 3630 hCoV-19/England/LIVE-A47BE/2020\|EPI_ISL_500043\|202 | ···················· | ···················· | ···················· | ···················· | ·······NNNNNNNNNNNNN | NNNNNNNNNNNNNNNNNNNN | 1 |
| F | 3632 hCoV-19/England/LIVE-A47FA/2020\|EPI_ISL_500045\|202 | ···················· | ···················· | ···················· | ···················· | ·······NNNNNNNNNNNNN | NNNNNNNNNNNNNNNNNNNN | 1 |
| F | 3633 hCoV-19/England/LIVE-A4815/2020\|EPI_ISL_500046\|202 | ···················· | ···················· | ···················· | ···················· | ·······NNNNNNNNNNNNN | ···················· | 1 |
| F | 3634 hCoV-19/England/LIVE-A4824/2020\|EPI_ISL_500047\|202 | ···················· | ···················· | ···················· | ···················· | ·······NNNNNNNNNNNNN | ···················· | 1 |
| F | 3636 hCoV-19/England/LIVE-A4860/2020\|EPI_ISL_500049\|202 | ···················· | ···················· | ···················· | ···················· | ·······NNNNNNNNNNNNN | NNNNNNNNNNNNNNNNNNNN | 1 |
| F | 3637 hCoV-19/France/PAC-41071320/2020\|EPI_ISL_505003\|20 | ···················· | ···················· | ···················· | ···················· | NNNNNNNNNNNNNNNNNNNN | NNNNNNNNNNNNNNNNNNNN | 1 |
| F | 3872 hCoV-19/India/UN-N7/2020\|EPI_ISL_497762\|2020 | ···················· | ···················· | ···················· | ···················· | NNNNNNNNNNNN········ | ···················· | 1 |
| F | 3903 hCoV-19/England/LIVE-A488E/2020\|EPI_ISL_500050\|202 | ···················· | ···················· | ···················· | ···················· | ·······NNNNNNNNNNNNN | NNNNNNNNNNNNNNNNNNNN | 1 |
| F | 3904 hCoV-19/England/LIVE-A48BB/2020\|EPI_ISL_500051\|202 | ···················· | ···················· | ···················· | ···················· | ·······NNNNNNNNNNNNN | NNNNNNNNNNNNNNNNNNNN | 1 |
| F | 3905 hCoV-19/England/LIVE-A48CA/2020\|EPI_ISL_500052\|202 | ···················· | ···················· | ···················· | ···················· | ·······NNNNNNNNNNNNN | NNNNNNNNNNNNNNNNNNNN | 1 |
| F | 3906 hCoV-19/England/LIVE-A48E8/2020\|EPI_ISL_500053\|202 | ···················· | ···················· | ···················· | ···················· | ·······NNNNNNNNNNNNN | NNNNNNNNNNNNNNNNNNNN | 1 |
| F | 3907 hCoV-19/England/LIVE-A48F7/2020\|EPI_ISL_500054\|202 | ···················· | ···················· | ···················· | ···················· | ·······NNNNNNNNNNNNN | NNNNNNNNNNNNNNNNNNNN | 1 |
| F | 3909 hCoV-19/England/LIVE-A495E/2020\|EPI_ISL_500056\|202 | ···················· | ···················· | ···················· | ···················· | ·······NNNNNNNNNNNNN | NNNNNNNNNNNNNNNNNNNN | 1 |
| F | 3912 hCoV-19/England/LIVE-A49A9/2020\|EPI_ISL_500059\|202 | ···················· | ···················· | ···················· | ···················· | ·······NNNNNNNNNNNNN | NNNNNNNNNNNNNNNNNNNN | 1 |
| F | 3913 hCoV-19/England/LIVE-A49E5/2020\|EPI_ISL_500060\|202 | ···················· | ···················· | ···················· | ···················· | ·······NNNNNNNNNNNNN | NNNNNNNNNNNNNNNNNNNN | 1 |
| F | 3914 hCoV-19/England/LIVE-A49F4/2020\|EPI_ISL_500061\|202 | ···················· | ···················· | ···················· | ···················· | ·······NNNNNNNNNNNNN | NNNNNNNNNNNNNNNNNNNN | 1 |
| F | 3915 hCoV-19/England/LIVE-A4A1F/2020\|EPI_ISL_500062\|202 | ···················· | ···················· | ···················· | ···················· | ·······NNNNNNNNNNNNN | NNNNNNNNNNNNNNNNNNNN | 1 |
| F | 3916 hCoV-19/England/LIVE-A4A2E/2020\|EPI_ISL_500063\|202 | ···················· | ···················· | ···················· | ···················· | ·······NNNNNNNNNNNNN | NNNNNNNNNNNNNNNNN··· | 1 |

```
F   3918 hCoV-19/England/LIVE-A4AC4/2020|EPI_ISL_500069|202   ····················  ····················  ····················  ····················  ·······NNNNNNNNNNNNN  NNNNNNNNNNNNNNNNNNNN  1
F   3919 hCoV-19/England/LIVE-A4AD3/2020|EPI_ISL_500070|202   ····················  ····················  ····················  ····················  ·······NNNNNNNNNNNNN  ····················  1
F   3923 hCoV-19/England/LIVE-A4B49/2020|EPI_ISL_500074|202   ····················  ····················  ····················  ····················  ·······NNNNNNNNNNNNN  NNNNNNNNNNNNNNNNNNNN  1
F   3924 hCoV-19/England/LIVE-A4B76/2020|EPI_ISL_500075|202   ····················  ····················  ····················  ····················  ·······NNNNNNNNNNNNN  ····················  1
F   3925 hCoV-19/England/LIVE-A4BC1/2020|EPI_ISL_500076|202   ····················  ····················  ····················  ····················  ·······NNNNNNNNNNNNN  ····················  1
F   3926 hCoV-19/England/LIVE-A4BEF/2020|EPI_ISL_500077|202   ····················  ·············A······  ····················  ····················  ·······NNNNNNNNNNNNN  NNNNNNNNNNNNNNNNNNNN  1
F   3927 hCoV-19/England/LIVE-A4C19/2020|EPI_ISL_500078|202   ····················  ····················  ····················  ····················  ·······NNNNNNNNNNNNN  ····················  1
F   3928 hCoV-19/England/LIVE-A4C55/2020|EPI_ISL_500079|202   ····················  ····················  ····················  ····················  ·······NNNNNNNNNNNNN  NNNNNNNNNNNNNNNNNNNN  1
F   3929 hCoV-19/England/LIVE-A4C73/2020|EPI_ISL_500080|202   ····················  ····················  ····················  ····················  ·······NNNNNNNNNNNNN  ····················  1
F   3930 hCoV-19/England/LIVE-A4CA0/2020|EPI_ISL_500081|202   ····················  ····················  ····················  ····················  ·······NNNNNNNNNNNNN  NN··················  1
F   3931 hCoV-19/England/LIVE-A4CCE/2020|EPI_ISL_500082|202   ····················  ····················  ····················  ····················  ·······NNNNNNNNNNNNN  NNNNNNNNNNNNNNNNNNNN  1
F   3943 hCoV-19/England/LIVE-A4E8C/2020|EPI_ISL_500094|202   ····················  ····················  ····················  ····················  ·······NNNNNNNNNNNNN  NNNNNNNNNNNNNNNNNNNN  1
F   3944 hCoV-19/England/LIVE-A4EC8/2020|EPI_ISL_500095|202   ····················  ····················  ····················  ····················  ·······NNNNNNNNNNNNN  ····················  1
F   3947 hCoV-19/England/LIVE-A4F5C/2020|EPI_ISL_500098|202   ····················  ····················  ····················  ····················  ·······NNNNNNNNNNNNN  ····················  1
F   3958 hCoV-19/USA/FL-BPHL-0632/2020|EPI_ISL_508836|2020-   ····················  ····················  ····················  ····················  NNNNNNNNNNNN········  ····················  1
F   4211 hCoV-19/England/LIVE-A4A79/2020|EPI_ISL_500065|202   ····················  ····················  ····················  ····················  ·······NNNNNNNNNNNNN  ····················  1
F   4212 hCoV-19/England/LIVE-A4A88/2020|EPI_ISL_500066|202   ····················  ····················  ····················  ····················  ·······NNNNNNNNNNNNN  ····················  1
F   4220 hCoV-19/USA/MI-MDHHS-SC21273/2020|EPI_ISL_507528|2   ····················  ····················  ····················  ····················  NNNNNNNNNNNNNNNNNNNN  NNNNNNNNNNNNNNNNNNNN  1
F   4675 hCoV-19/USA/MI-MDHHS-SC21174/2020|EPI_ISL_507431|2   ····················  ····················  ····················  ····················  NNNNNNNNNNNNNNNNNNNN  NNNNNNNNNNNNNNNNNNNN  1
F   4676 hCoV-19/USA/MI-MDHHS-SC21177/2020|EPI_ISL_507434|2   ····················  ····················  ····················  ····················  NNNNNNNNNNNNNNNNNNNN  NNNNNNNNNNNNNNNNNNNN  1
F   4677 hCoV-19/USA/MI-MDHHS-SC21178/2020|EPI_ISL_507435|2   ····················  ····················  ····················  ····················  NNNNNNNNNNNNNNNNNNNN  NNNNNNNNNNNNNNNNNNNN  1
F   4678 hCoV-19/USA/MI-MDHHS-SC21180/2020|EPI_ISL_507437|2   ····················  ····················  ····················  ····················  NNNNNNNNNNNNNNNNNNNN  NNNNNNNNNNNNNNNNNNNN  1
F   4679 hCoV-19/USA/MI-MDHHS-SC21181/2020|EPI_ISL_507438|2   ····················  ····················  ····················  ····················  NNNNNNNNNNNNNNNNNNNN  NNNNNNNNNNNNNNNNNNNN  1
F   4680 hCoV-19/USA/MI-MDHHS-SC21184/2020|EPI_ISL_507441|2   ····················  ····················  ····················  ····················  NNNNNNNNNNNNNNNNNNNN  NNNNNNNNNNNNNNNNNNNN  1
F   4681 hCoV-19/USA/MI-MDHHS-SC21185/2020|EPI_ISL_507442|2   ····················  ····················  ····················  ····················  NNNNNNNNNNNNNNNNNNNN  NNNNNNNNNNNNNNNNNNNN  1
F   4682 hCoV-19/USA/MI-MDHHS-SC21187/2020|EPI_ISL_507444|2   ····················  ····················  ····················  ····················  NNNNNNNNNNNNNNNNNNNN  NNNNNNNNNNNNNNNNNNNN  1
F   4684 hCoV-19/USA/MI-MDHHS-SC21189/2020|EPI_ISL_507446|2   ····················  ····················  ····················  ····················  NNNNNNNNNNNNNNNNNNNN  ····················  1
F   4685 hCoV-19/USA/MI-MDHHS-SC21202/2020|EPI_ISL_507459|2   ····················  ····················  ····················  ····················  NNNNNNNNNNNNNNNNNNNN  NNNNNNNNNNNNNNNNNNNN  1
F   4686 hCoV-19/USA/MI-MDHHS-SC21203/2020|EPI_ISL_507460|2   ····················  ····················  ····················  ····················  NNNNNNNNNNNNNNNNNNNN  NNNNNNNNNNNNNNNNNNNN  1
F   4687 hCoV-19/USA/MI-MDHHS-SC21205/2020|EPI_ISL_507462|2   ····················  ····················  ····················  ····················  NNNNNNNNNNNNNNNNNNNN  NNNNNNNNNNNNNNNNNNNN  1
F   4688 hCoV-19/USA/MI-MDHHS-SC21207/2020|EPI_ISL_507464|2   ····················  ····················  ····················  ····················  NNNNNNNNNNNNNNNNNNNN  NNNNNNNNNNNNNNNNNNNN  1
F   4689 hCoV-19/USA/MI-MDHHS-SC21208/2020|EPI_ISL_507465|2   ····················  ····················  ····················  ····················  NNNNNNNNNNNNNNNNNNNN  NNNNNNNNNNNNNNNNNNNN  1
F   4690 hCoV-19/USA/MI-MDHHS-SC21209/2020|EPI_ISL_507466|2   ····················  ····················  ····················  ····················  NNNNNNNNNNNNNNNNNNNN  NNNNNNNNNNNNNNNNNNNN  1
F   4691 hCoV-19/USA/MI-MDHHS-SC21210/2020|EPI_ISL_507467|2   ····················  ····················  ····················  ····················  RNNNNNNNNNNNNNNNNNNN  NNNNNNNNNNNNNNNNNNNN  1
F   4692 hCoV-19/USA/MI-MDHHS-SC21211/2020|EPI_ISL_507468|2   ····················  ····················  ····················  ····················  NNNNNNNNNNNNNNNNNNNN  NNNNNNNNNNNNNNNNNNNN  1
F   4693 hCoV-19/USA/MI-MDHHS-SC21212/2020|EPI_ISL_507469|2   ····················  ····················  ····················  ····················  NNNNNNNNNNNNNNNNNNNN  NNNNNNNNNNNNNNNNNNNN  1
F   4694 hCoV-19/USA/MI-MDHHS-SC21213/2020|EPI_ISL_507470|2   ····················  ····················  ····················  ····················  NNNNNNNNNNNNNNNNNNNN  NNNNNNNNNNNNNNNNNNNN  1
F   4695 hCoV-19/USA/MI-MDHHS-SC21214/2020|EPI_ISL_507471|2   ····················  ····················  ····················  ····················  NNNNNNNNNNNNNNNNNNNN  NNNNNNNNNNNNNNNNNNNN  1
F   4705 hCoV-19/USA/FL-BPHL-0510/2020|EPI_ISL_508713|2020-   ····················  ····················  ····················  ····················  NNNNNNNNNNNNNNNNNNNN  ····················  1
F   4710 hCoV-19/USA/FL-BPHL-0517/2020|EPI_ISL_508720|2020-   ····················  ····················  ····················  ····················  NNNNNNNNNNNN········  ····················  1
F   4750 hCoV-19/USA/FL-BPHL-0570/2020|EPI_ISL_508774|2020-   ····················  ····················  ····················  ····················  ········NNNNNNNNNNN·  ····················  1
F   4752 hCoV-19/USA/FL-BPHL-0572/2020|EPI_ISL_508776|2020-   ····················  ····················  ····················  ····················  NNNNNNNNNNNNNNNNNNNN  NNNNNNNNNNNNNNNNNNNN  1
F   4755 hCoV-19/USA/FL-BPHL-0575/2020|EPI_ISL_508779|2020-   ····················  ····················  ····················  ····················  NNNNNNNNNNNNNNNNNNNN  ··············NNNNNN  1
F   4824 hCoV-19/Wales/PHWC-169A51/2020|EPI_ISL_514570|2020   ····················  ····················  ····················  ····················  NNNNNNNNNNNNNNNNNNNN  ····················  1
F   4828 hCoV-19/Wales/PHWC-169A9D/2020|EPI_ISL_514574|2020   ····················  ····················  ····················  ····················  ···········NNNNNNNNN  NNNNNNNNNNNNNNNNNNNN  1
F   4829 hCoV-19/Wales/PHWC-169AAC/2020|EPI_ISL_514575|2020   ····················  ····················  ····················  ····················  ·NNNNNNNNNNNNNNNNNNN  ····················  1
F   5283 hCoV-19/USA/WI-UW-930/2020|EPI_ISL_509930|2020-07-   ····················  ····················  ····················  ····················  NNNNNNNNNNNNNNNNNNNN  NNNNNNNNNNNNNNNNNNNN  1
F   5322 hCoV-19/SaudiArabia/KAUST-MADINAH691/2020|EPI_ISL_   ····················  ····················  ····················  ····················  ·······NNNNNNNNNNNNN  NNNNNNNNNNNNNNNNNNNN  1
F   5412 hCoV-19/Spain/CT-IBV-005574/2020|EPI_ISL_508627|20   ····················  ····················  ····················  ····················  NNNNNNNNNNNNNNNNNNNN  NNNNNNNNNNNNNNNNNNNN  1
F   5413 hCoV-19/Spain/CT-IBV-005590/2020|EPI_ISL_508639|20   ····················  ····················  ····················  ····················  NNNNNNNNNNNNNNNNNNNN  ····················  1
F   5414 hCoV-19/Spain/CT-IBV-005591/2020|EPI_ISL_508640|20   ····················  ····················  ····················  ····················  NNNNNNNNNNNNNNNNNNNN  NNNNNNNNNNNNNNNNNNNN  1
F   5415 hCoV-19/Spain/CT-IBV-005593/2020|EPI_ISL_508641|20   ····················  ····················  ····················  ····················  NNNNNNNNNNNNNNNNNNNN  NNNNNNNNNNNNNNNNNNNN  1
F   5416 hCoV-19/Spain/CT-IBV-005594/2020|EPI_ISL_508642|20   ····················  ····················  ····················  ····················  NNNNNNNNNNNNNNNNNNNN  NNNNNNNNNNNNNNNNNNNN  1
F   5417 hCoV-19/Spain/CT-IBV-005595/2020|EPI_ISL_508643|20   ····················  ····················  ····················  ····················  NNNNNNNNNNNNNNNNNNNN  NNNNNNNNNNNNNNNNNNNN  1
F   5418 hCoV-19/Spain/CT-IBV-005599/2020|EPI_ISL_508646|20   ····················  ····················  ····················  ····················  NNNNNNNNNNNNNNNNNNNN  NNNNNNNNNNNNNNNNNNNN  1
F   5419 hCoV-19/Spain/CT-IBV-005601/2020|EPI_ISL_508647|20   ····················  ····················  ····················  ····················  NNNNNNNNNNNNNNNNNNNN  NNNNNNNNNNNNNNNNNNNN  1
F   5420 hCoV-19/Spain/CT-IBV-005608/2020|EPI_ISL_508653|20   ····················  ····················  ····················  ····················  NNNNNNNNNNNNNNNNNNNN  NNNNNNNNNNNNNNNNNNNN  1
F   5422 hCoV-19/Spain/CT-IBV-005630/2020|EPI_ISL_508663|20   ····················  ····················  ····················  ····················  NNNNNNNNNNNNNNNNNNNN  NNNNNNNNNNNNNNNNNNNN  1
F   5423 hCoV-19/Spain/CT-IBV-005634/2020|EPI_ISL_508667|20   ····················  ····················  ····················  ····················  ·NNNNNNNNNNN········  ····················  1
F   5424 hCoV-19/Spain/CT-IBV-005635/2020|EPI_ISL_508676|20   ····················  ····················  ····················  ····················  NNNNNNNNNNNNNNNNNNNN  ····················  1
F   5429 hCoV-19/USA/WI-UW-943/2020|EPI_ISL_509943|2020-05-   ····················  ····················  ····················  ····················  NNNNNNNNNNNNNNNNNNNN  NNNNNNNNNNNNNNNNNNNN  1
F   5472 hCoV-19/USA/WI-UW-992/2020|EPI_ISL_509987|2020-07-   ····················  ····················  ····················  ····················  NNNNNNNNNNNNNNNNNNNN  NNNNNNNNNNNNNNNNNNNN  1
F   5490 hCoV-19/Portugal/PT0672/2020|EPI_ISL_510931|2020-0   ····················  ····················  ····················  ····················  NNNNNNNNNNNNNNNNNNNN  NNNNNNNNNNNNNNNNNNNN  1
F   5505 hCoV-19/Portugal/PT0687/2020|EPI_ISL_510946|2020-0   ····················  ····················  ····················  ····················  NNNNNNNNNNNNNNNNNNNN  ····················  1
F   5632 hCoV-19/South Africa/KRISP-K002443/2020|EPI_ISL_51   ····················  ····················  ····················  ····················  ····················  NNNNN···············  1
F   5701 hCoV-19/Ireland/D-NVRL-71IRL21349/2020|EPI_ISL_500   ····················  ····················  ····················  ····················  NNNNNNNNNNNNNNNNNNNN  NNNNNNNNNNNNNNNNNNNN  1
F   5726 hCoV-19/Portugal/PT1253/2020|EPI_ISL_511214|2020-0   ····················  ····················  ····················  ····················  NNNNNNNNNNNNNNNNNNNN  ····················  1
F   5734 hCoV-19/Portugal/PT1283/2020|EPI_ISL_511244|2020-0   ····················  ····················  ····················  ····················  NNNNNNNNNNNNNNNNNNNN  ····················  1
F   5737 hCoV-19/Portugal/PT1286/2020|EPI_ISL_511247|2020-0   ····················  ····················  ····················  ····················  NNNNNNNNNNNNNNNNNNNN  ····················  1
F   5746 hCoV-19/Portugal/PT1296/2020|EPI_ISL_511256|2020-0   ····················  ····················  ····················  ····················  NNNNNNNNNNNNNNNNNNNN  ····················  1
F   5774 hCoV-19/Wales/PHWC-1696E4/2020|EPI_ISL_512500|2020   ····················  ····················  ····················  ····················  ········NNNNNNNNNNNN  NNNNNNNNNNNNNNNNNNNN  1
F   5776 hCoV-19/Wales/PHWC-16970F/2020|EPI_ISL_512502|2020   ····················  ····················  ····················  ····················  NNNN················  ····················  1
F   5810 hCoV-19/USA/FL-BPHL-0753/2020|EPI_ISL_512559|2020-   ····················  ····················  ····················  ····················  NNNN················  ····················  1
F   5817 hCoV-19/USA/FL-BPHL-0775/2020|EPI_ISL_512579|2020-   ····················  ····················  ····················  ····················  NNNNNNNNNNNNNNNNNNNN  NNNNNNNNNNNNNNNNNNNN  1
F   5846 hCoV-19/USA/CA-CZB-2419/2020|EPI_ISL_513840|2020-0   ····················  ····················  ····················  ····················  ·······NNNNNNNNNNNNN  NNNNNNNNNNNNNNNNNNNN  1
F   5847 hCoV-19/USA/CA-CZB-2420/2020|EPI_ISL_513841|2020-0   ····················  ····················  ····················  ····················  NNNN················  ····················  1
F   5893 hCoV-19/Spain/RI-IBV-005974/2020|EPI_ISL_500408|20   ····················  ····················  ····················  ····················  NNNNNNNNNNNNNNNNNNNN  NNNNNNNNNNNNNNNNNNNN  1
F   5894 hCoV-19/Spain/RI-IBV-005975/2020|EPI_ISL_500409|20   ····················  ····················  ····················  ····················  ·············NNNNNNN  NNNNNNNNNNNNNNN·····  1
F   5895 hCoV-19/Spain/RI-IBV-005976/2020|EPI_ISL_500410|20   ····················  ····················  ····················  ····················  NNNN················  ····················  1
F   5897 hCoV-19/Spain/RI-IBV-005978/2020|EPI_ISL_500412|20   ····················  ····················  ····················  ····················  NNNNNNNNNNNNNNNNNNNN  NNNNNNNNNNNNNNNNNNNN  1
```

| | | | | | | | | |
|---|---|---|---|---|---|---|---|---|
| F | 5899 hCoV-19/Spain/RI-IBV-005982/2020\|EPI_ISL_500416\|20 | ···················· | ···················· | ···················· | ···················· | NNNNNNNNNNNNNNNNNNNN | NNNNNNNNNNNNNNNNNNNN | 1 |
| F | 5900 hCoV-19/Spain/RI-IBV-005983/2020\|EPI_ISL_500417\|20 | ···················· | ···················· | ···················· | ···················· | NNNNNNNNNNNNNNNNNNNN | NNNNNNNNNNNNNNNNNNNN | 1 |
| F | 5901 hCoV-19/Spain/RI-IBV-005984/2020\|EPI_ISL_500418\|20 | ···················· | ···················· | ···················· | ···················· | NNNNNNNNNNNNNNNNNNNN | NNNNNNNNNNNNNNNNNNNN | 1 |
| F | 5904 hCoV-19/Spain/RI-IBV-005993/2020\|EPI_ISL_500425\|20 | ···················· | ···················· | ···················· | ···················· | NNNNNNN············· | ···················· | 1 |
| F | 5911 hCoV-19/Spain/RI-IBV-006008/2020\|EPI_ISL_500439\|20 | ···················· | ···················· | ···················· | ···················· | NNNNNNNNNNNNNNNNNNNN | NNNNNNNNNNNNNNNNNNNN | 1 |
| F | 5912 hCoV-19/Spain/RI-IBV-006015/2020\|EPI_ISL_500445\|20 | ···················· | ···················· | ···················· | ···················· | NNNNNNNNNNNNNNNNNNNN | NNNNNNNNNNNNNNNNNNNN | 1 |
| F | 6003 hCoV-19/Portugal/PT1260/2020\|EPI_ISL_511221\|2020-0 | ···················· | ···················· | ···················· | ···················· | NNNNN··············· | ···················· | 1 |
| F | 6029 hCoV-19/Scotland/CVR3928/2020\|EPI_ISL_512471\|2020- | ···················· | ···················· | ···················· | ···················· | NNNNNNNNNNNNNNNNNNNN | NNNNNNNNNNNNNNNNNNNN | 1 |
| F | 6038 hCoV-19/Wales/PHWC-1695C9/2020\|EPI_ISL_512482\|2020 | ···················· | ···················· | ···················· | ···················· | NNNNNNNNNNNNNNNNNNNN | NNNNNNN············· | 1 |
| F | 6041 hCoV-19/Wales/PHWC-1695F6/2020\|EPI_ISL_512485\|2020 | ···················· | ···················· | ···················· | ···················· | NNNNNNNNNNN········· | ···················· | 1 |
| F | 6048 hCoV-19/Wales/PHWC-16966C/2020\|EPI_ISL_512492\|2020 | ···················· | ···················· | ···················· | ···················· | NNNNNNNNNNNNNNNNNNNN | ···················· | 1 |
| F | 6058 hCoV-19/USA/FL-BPHL-0744/2020\|EPI_ISL_512550\|2020- | ···················· | ···················· | ···················· | ···················· | NNNNNNNNNNNNNNNNNNNN | ···················· | 1 |
| F | 6060 hCoV-19/USA/FL-BPHL-0748/2020\|EPI_ISL_512554\|2020- | ···················· | ···················· | ···················· | ···················· | NNNNNNNNNNNNNNNNNNNN | NNNNNNNNNNN·NNNNNNNN | 1 |
| F | 6126 hCoV-19/USA/CA-CZB-2498/2020\|EPI_ISL_513885\|2020-0 | ···················· | ·A·················· | ···················· | ···················· | NNNNNNNNNNNNNNNNNNNN | ···················· | 1 |
| F | 6146 hCoV-19/Spain/PV-IBV-004240/2020\|EPI_ISL_500304\|20 | ···················· | ···················· | ···················· | ···················· | NNNNNNNNNNNNNNNNNNNN | NNNNNNNNNNNNNNNNNNNN | 1 |
| F | 6147 hCoV-19/Spain/PV-IBV-004241/2020\|EPI_ISL_500305\|20 | ···················· | ···················· | ···················· | ···················· | NNNNNNNNNNNNNNNNNNNN | NNNNNNNNNNNNNNNNNNNN | 1 |
| F | 6149 hCoV-19/Spain/PV-IBV-004243/2020\|EPI_ISL_500307\|20 | ···················· | ···················· | ···················· | ···················· | NNNNNNNNNNNNNNNNNNNN | NNNNNNNNNNNNNNNNNNNN | 1 |
| F | 6150 hCoV-19/Spain/PV-IBV-004244/2020\|EPI_ISL_500308\|20 | ···················· | ···················· | ···················· | ···················· | NNNNNNNNNNNNNNNNNNNN | NNNNNNNNNNNNNNNNNNNN | 1 |
| F | 6151 hCoV-19/Spain/PV-IBV-004245/2020\|EPI_ISL_500309\|20 | ···················· | ···················· | ···················· | ···················· | NNNNNNNNNNNNNNNNNNNN | NNNNNNNNNNNNNNNNNNNN | 1 |
| F | 6152 hCoV-19/Spain/PV-IBV-004247/2020\|EPI_ISL_500311\|20 | ···················· | ···················· | ···················· | ···················· | NNNNNNNNNNNNNNNNNNNN | NNNNNNNNNNNNNNNNNNNN | 1 |
| F | 6153 hCoV-19/Spain/RI-IBV-006019/2020\|EPI_ISL_500449\|20 | ···················· | ···················· | ···················· | ···················· | NNNNNNN············· | ···················· | 1 |
| F | 6154 hCoV-19/Spain/RI-IBV-006020/2020\|EPI_ISL_500450\|20 | ···················· | ···················· | ···················· | ···················· | NNNNNNN············· | ···················· | 1 |
| F | 6155 hCoV-19/Spain/RI-IBV-006021/2020\|EPI_ISL_500451\|20 | ···················· | ···················· | ···················· | ···················· | NNNNNNNNNNNNNNNNNNNN | NNNNNNNNNNNNNNNNNNNN | 1 |
| F | 6156 hCoV-19/Spain/RI-IBV-006024/2020\|EPI_ISL_500454\|20 | ···················· | ···················· | ···················· | ···················· | NNNNNNNNNNNNNNNNNNNN | NNNNNNNNNNNNNNNNNNNN | 1 |
| F | 6157 hCoV-19/Spain/RI-IBV-006025/2020\|EPI_ISL_500455\|20 | ···················· | ···················· | ···················· | ···················· | NNNNNNNNNNNNNNNNNNNN | NNNNNNNNNNNNNNNNNNNN | 1 |
| F | 6158 hCoV-19/Spain/RI-IBV-006026/2020\|EPI_ISL_500456\|20 | ···················· | ···················· | ···················· | ···················· | ·NNNNNNNN··········· | ···················· | 1 |
| F | 6165 hCoV-19/Brazil/PE-IAM215/2020\|EPI_ISL_500471\|2020- | ···················· | ···················· | ···················· | ···················· | NNNNNNNNNNNNNNNNN··· | ···················· | 1 |
| F | 6217 hCoV-19/USA/MN-MDH-1368/2020\|EPI_ISL_507956\|2020-0 | ···················· | ···················· | ···················· | ···················· | ··NNNNNNNNNN········ | ···················· | 1 |
| F | 6236 hCoV-19/USA/MN-MDH-1388/2020\|EPI_ISL_507975\|2020-0 | ···················· | ···················· | ···················· | ···················· | ·NNNNN·············· | ···················· | 1 |
| F | 6300 hCoV-19/Portugal/PT1233/2020\|EPI_ISL_511194\|2020-0 | ···················· | ···················· | ···················· | ···················· | NN··NN·············· | ···················· | 1 |
| F | 6315 hCoV-19/Scotland/CVR1535/2020\|EPI_ISL_512438\|2020- | ···················· | ···················· | ···················· | ···················· | NNNNNNNNNNNNNNNNNNNN | NNNNNNNNNNNN········ | 1 |
| F | 6319 hCoV-19/Scotland/CVR1662/2020\|EPI_ISL_512446\|2020- | ···················· | ···················A | ···················· | ···················· | ·······NNNNNNNNNNNNN | NNNNNNNNNNNNNNNNNNNN | 1 |
| F | 6321 hCoV-19/Scotland/CVR1708/2020\|EPI_ISL_512448\|2020- | ···················· | ···················· | ···················· | ···················· | ·······NNNNNNNNNNNNN | ···················· | 1 |
| F | 6322 hCoV-19/Scotland/CVR1711/2020\|EPI_ISL_512449\|2020- | ···················· | ···················· | ···················· | ···················· | ·······NNNNNNNNNNNNN | NNNNNNNNNNNNNNNNNNNN | 1 |
| F | 6323 hCoV-19/Scotland/CVR1857/2020\|EPI_ISL_512452\|2020- | ···················· | ···················· | ···················· | ···················· | NNNNNNNNNNNNNNNNNNNN | ···················· | 1 |
| F | 6324 hCoV-19/Scotland/CVR1862/2020\|EPI_ISL_512453\|2020- | ···················· | ···················· | ···················· | ···················· | ·······NNNNNNNNNNNNN | ···················· | 1 |
| F | 6325 hCoV-19/Scotland/CVR1864/2020\|EPI_ISL_512454\|2020- | ···················· | ···················· | ···················· | ···················· | NNNNNNNNNNNNNNNNNNNN | ···················· | 1 |
| F | 6327 hCoV-19/Scotland/CVR3905/2020\|EPI_ISL_512456\|2020- | ···················· | ···················· | ···················· | ···················· | NNNNNNNNNNNNNNNNNNNN | ···················· | 1 |
| F | 6328 hCoV-19/Scotland/CVR3906/2020\|EPI_ISL_512457\|2020- | ···················· | ···················· | ···················· | ···················· | NNNNNNNNNNNNNNNNNNNN | NNNNNNNNNNNNNNNNNNNN | 1 |
| F | 6329 hCoV-19/Scotland/CVR3907/2020\|EPI_ISL_512458\|2020- | ···················· | ···················· | ···················· | ···················· | NNNNNNNNNNNNNNNNNNNN | ···················· | 1 |
| F | 6333 hCoV-19/Scotland/CVR3914/2020\|EPI_ISL_512462\|2020- | ···················· | ···················· | ···················· | ···················· | NNNNNNNNNNNNNNNNNNNN | ···················· | 1 |
| F | 6335 hCoV-19/Scotland/CVR3919/2020\|EPI_ISL_512467\|2020- | ···················· | ···················· | ···················· | ···················· | NNNNNNNNNNNNNNNNNNNN | ···················· | 1 |
| F | 6337 hCoV-19/Scotland/CVR3922/2020\|EPI_ISL_512469\|2020- | ···················· | ···················· | ···················· | ···················· | NNNNNNNNNNNNNNNNNNNN | ···················· | 1 |
| F | 6338 hCoV-19/Scotland/CVR3933/2020\|EPI_ISL_512475\|2020- | ···················· | ···················· | ···················· | ···················· | NNNNNNNNNNNNNNNNNNNN | ···················· | 1 |
| F | 6413 hCoV-19/India/MH-NCCS-NR2107/2020\|EPI_ISL_496569\|2 | ···················· | ···················· | ···················· | ···················· | ········NNNNNNNNNNNN | ···················· | 1 |
| F | 6419 hCoV-19/Spain/PV-IBV-004236/2020\|EPI_ISL_500300\|20 | ···················· | ···················· | ···················· | ···················· | ·NNNNNNNNNNNNNNNNNNN | NNNNNNNN············ | 1 |
| F | 6420 hCoV-19/Spain/PV-IBV-004237/2020\|EPI_ISL_500301\|20 | ···················· | ···················· | ···················· | ···················· | NNNNNNNNNNNNNNNNNNNN | NNNNNNNNNNNNNNNNNNNN | 1 |
| F | 6421 hCoV-19/Spain/PV-IBV-004238/2020\|EPI_ISL_500302\|20 | ···················· | ···················· | ···················· | ···················· | NNNNNNNNNNNNNNNNNNNN | NNNNN·········N····· | 1 |
| F | 6422 hCoV-19/Spain/PV-IBV-004239/2020\|EPI_ISL_500303\|20 | ···················· | ···················· | ···················· | ···················· | NNNNNNNNNNNNNNNNNNNN | NNNNNNNNNNNNNNNNNNNN | 1 |
| F | 6423 hCoV-19/Spain/PV-IBV-004251/2020\|EPI_ISL_500315\|20 | ···················· | ···················· | ···················· | ···················· | NNNNNNNNNNNNNNNNNNNN | ·············NN····· | 1 |
| F | 6424 hCoV-19/Spain/PV-IBV-004255/2020\|EPI_ISL_500319\|20 | ···················· | ···················· | ···················· | ···················· | NNNNNNNNNNNNNNNNNNNN | NNNNNNNNNNNNNNNNNNNN | 1 |
| F | 6425 hCoV-19/Spain/PV-IBV-004256/2020\|EPI_ISL_500320\|20 | ···················· | ···················· | ···················· | ···················· | NNNNNNNNNNNNNNNNNNNN | NNNNNNNNNNNNNNNNNNNN | 1 |
| F | 6426 hCoV-19/Spain/PV-IBV-004257/2020\|EPI_ISL_500321\|20 | ···················· | ···················· | ···················· | ···················· | NNNNNNNNNNNNNNNNNNNN | NNNNNNNNNNNNNNNNNNNN | 1 |
| F | 6427 hCoV-19/Spain/PV-IBV-004258/2020\|EPI_ISL_500322\|20 | ···················· | ···················· | ···················· | ···················· | ·NNNNNNNNNNNNNNNNNNN | NNNNNNNNNN·········· | 1 |
| F | 6428 hCoV-19/Spain/PV-IBV-004259/2020\|EPI_ISL_500323\|20 | ···················· | ···················· | ···················· | ···················· | NNNNNNNNNNNNNNNNNNNN | NNNNNNNNNNNNNNNNNNNN | 1 |
| F | 6429 hCoV-19/Spain/PV-IBV-004260/2020\|EPI_ISL_500324\|20 | ···················· | ···················· | ···················· | ···················· | NNNNNNNNNNNNNNNNNNNN | NNNNNNNNNNNNNNNNNNNN | 1 |
| F | 6430 hCoV-19/Spain/PV-IBV-004261/2020\|EPI_ISL_500325\|20 | ···················· | ···················· | ···················· | ···················· | NNNNNNNNNNNNNNNNNNNN | NNNNNNNNNNNNNNNNNNNN | 1 |
| F | 6431 hCoV-19/Spain/AR-IBV-005065/2020\|EPI_ISL_500349\|20 | ···················· | ···················· | ···················· | ···················· | NNNNNNNNNNNNNNNNNNNN | NNNNNNNNNNNNNNNNNNNN | 1 |
| F | 6432 hCoV-19/Spain/AR-IBV-005092/2020\|EPI_ISL_500354\|20 | ···················· | ···················· | ···················· | ···················· | NNNNNNNNNNNNNNNNNNNN | NNNNNNNNNNNNNNNNNNNN | 1 |
| F | 6434 hCoV-19/Spain/RI-IBV-005936/2020\|EPI_ISL_500372\|20 | ···················· | ···················· | ···················· | ···················· | NNNNNNNNNNNNNNNNNNNN | NNNNNNNNNNNNNNN····· | 1 |
| F | 6436 hCoV-19/Spain/RI-IBV-005941/2020\|EPI_ISL_500376\|20 | ···················· | ···················· | ···················· | ···················· | NNNNNNNNNNNNNNNNNNNN | NNNNNNNNNNNNNNN····· | 1 |
| F | 6437 hCoV-19/Spain/RI-IBV-005945/2020\|EPI_ISL_500379\|20 | ···················· | ···················· | ···················· | ···················· | NNNNNNNNNNNNNNNNNNNN | ··········NNNNN····· | 1 |
| F | 6538 hCoV-19/DRC/6392/2020\|EPI_ISL_513616\|2020-05-04 | ···················· | ···················· | ···················· | ···················· | NNNNNNN············· | ···················· | 1 |
| F | 6771 hCoV-19/Spain/PV-IBV-004262/2020\|EPI_ISL_500326\|20 | ···················· | ···················· | ···················· | ···················· | NNNNNNNNNNNNNNNNNNNN | NNNNNNNNNNNNNNNNNNNN | 1 |
| F | 6772 hCoV-19/Spain/PV-IBV-004264/2020\|EPI_ISL_500328\|20 | ···················· | ···················· | ···················· | ···················· | NNNNNNNNNNNNNNNNNNNN | NNNNNNNNNNNNNNNNNNNN | 1 |
| F | 6773 hCoV-19/Spain/PV-IBV-004265/2020\|EPI_ISL_500329\|20 | ···················· | ···················· | ···················· | ···················· | NNNNNNNNNNNNNNNNNNNN | NNNNNNNNNNNNNNNNNNNN | 1 |
| F | 6774 hCoV-19/Spain/PV-IBV-004267/2020\|EPI_ISL_500331\|20 | ···················· | ···················· | ···················· | ···················· | NNNNNNNNNNNNNNNNNNNN | NNNNNNNNNNNNNNNNNNNN | 1 |
| F | 6775 hCoV-19/Spain/PV-IBV-004268/2020\|EPI_ISL_500332\|20 | ···················· | ···················· | ···················· | ···················· | NNNNNNNNNNNNNNNNNNNN | NNNNNNNNNNNNNNNNNNNN | 1 |
| F | 6776 hCoV-19/Spain/PV-IBV-004269/2020\|EPI_ISL_500333\|20 | ···················· | ···················· | ···················· | ···················· | NNNNNNNNNNNNNNNNNNNN | NNNNNNNNNNNNNNNNNNNN | 1 |
| F | 6777 hCoV-19/Spain/PV-IBV-004270/2020\|EPI_ISL_500334\|20 | ···················· | ···················· | ···················· | ···················· | NNNNNNNNNNNNNNNNNNNN | NNNNNNNNNNNNNNNNNNNN | 1 |
| F | 6778 hCoV-19/Spain/PV-IBV-004271/2020\|EPI_ISL_500335\|20 | ···················· | ···················· | ···················· | ···················· | NNNNNNNNNNNNNNNNNNNN | NNNNNNNNNNNNNNNNNNNN | 1 |
| F | 6779 hCoV-19/Spain/AR-IBV-005029/2020\|EPI_ISL_500337\|20 | ···················· | ···················· | ···················· | ···················· | NNNNNNNNNNNNNNNNNNNN | NNNNNNNNNNNNNNNNNNNN | 1 |
| F | 6780 hCoV-19/Spain/AR-IBV-005052/2020\|EPI_ISL_500344\|20 | ···················· | ···················· | ···················· | ···················· | NNNNNNNNNNNNNNNNNNNN | NNNNNNNNNNNNNNNNNNNN | 1 |
| F | 6781 hCoV-19/Spain/AR-IBV-005061/2020\|EPI_ISL_500347\|20 | ···················· | ···················· | ···················· | ···················· | NNNNNNNNNNNNNNNNNNNN | NNNNNNNNNNNNNNNNNNNN | 1 |
| F | 6917 hCoV-19/Spain/AR-IBV-005054/2020\|EPI_ISL_500287\|20 | ···················· | ···················· | ···················· | ···················· | NNNNNNNNNNNNNNNNNNNN | NNNNNNNNNNNNNNNNNNNN | 1 |
| F | 6950 hCoV-19/USA/MI-MDHHS-SC20954/2020\|EPI_ISL_507743\|2 | ···················· | ···················· | ···················· | ···················· | NNNNNNNNNNNNNNNNNNNN | NNNNNNNNNNNNNNNNNNNN | 1 |
| F | 6951 hCoV-19/USA/MI-MDHHS-SC20955/2020\|EPI_ISL_507744\|2 | ···················· | ···················· | ···················· | ···················· | NNNNNNNNNNNNNNNNNNNN | NNNNNNNNNNNNNNNNNNNN | 1 |
| F | 6952 hCoV-19/USA/MI-MDHHS-SC20956/2020\|EPI_ISL_507745\|2 | ···················· | ···················· | ···················· | ···················· | NNNNNNNNNNNNNNNNNNNN | NNNNNNNNNNNNNNNNNNNN | 1 |
| F | 6953 hCoV-19/USA/MI-MDHHS-SC20957/2020\|EPI_ISL_507746\|2 | ···················· | ···················· | ···················· | ···················· | NNNNNNNNNNNNNNNNNNNN | NNNNNNNNNNNNNNNNNNNN | 1 |
| F | 7021 hCoV-19/Brazil/RJ-INCA-C187/2020\|EPI_ISL_513522\|20 | ···················· | ···················· | ···················· | ···················· | NNNNNNNNNNNNNNNNNNNN | ···················· | 1 |

<table>
<tr><td>F</td><td>7214</td><td>hCoV-19/South Africa/KRISP-0569/2020|EPI_ISL_49810</td><td>····················</td><td>····················</td><td>····················</td><td>····················</td><td>·NNNNNNNNNNNNNNNNNNN</td><td>NNNNNNNNNNNNNNNNNNNN</td><td>1</td></tr>
<tr><td>F</td><td>7256</td><td>hCoV-19/Spain/PV-IBV-004228/2020|EPI_ISL_500292|20</td><td>····················</td><td>····················</td><td>····················</td><td>····················</td><td>NNNNNNNNNNNNNNNNNNNN</td><td>NNNNNNNNNNNNNNNNNNNN</td><td>1</td></tr>
<tr><td>F</td><td>7258</td><td>hCoV-19/Spain/PV-IBV-004232/2020|EPI_ISL_500296|20</td><td>····················</td><td>····················</td><td>····················</td><td>····················</td><td>NNNNNNNNNNNNNNNNNNNN</td><td>NNNNNNNN·NNNNNN·····</td><td>1</td></tr>
<tr><td>F</td><td>7259</td><td>hCoV-19/Spain/PV-IBV-004233/2020|EPI_ISL_500297|20</td><td>····················</td><td>····················</td><td>····················</td><td>····················</td><td>NNNNNNNNNNNNNNNNNNNN</td><td>NNNNNNNNNNNNNNNNNNNN</td><td>1</td></tr>
<tr><td>F</td><td>7260</td><td>hCoV-19/Spain/PV-IBV-004234/2020|EPI_ISL_500298|20</td><td>····················</td><td>····················</td><td>····················</td><td>····················</td><td>NNNNNNNNNNNNNNNNNNNN</td><td>NNNNNNNNNNNNNNNNNNNN</td><td>1</td></tr>
<tr><td>F</td><td>7282</td><td>hCoV-19/USA/MI-MDHHS-SC20950/2020|EPI_ISL_507739|2</td><td>····················</td><td>····················</td><td>····················</td><td>····················</td><td>NNNNNNNNNNNNNNNNNNNN</td><td>NNNNNNNNNNNNNNNNNNNN</td><td>1</td></tr>
<tr><td>F</td><td>7283</td><td>hCoV-19/USA/MI-MDHHS-SC20951/2020|EPI_ISL_507740|2</td><td>····················</td><td>····················</td><td>····················</td><td>····················</td><td>NNNNNNNNNNNNNNNNNNNN</td><td>NNNNNNNNNNNNNNNNNNNN</td><td>1</td></tr>
<tr><td>F</td><td>7284</td><td>hCoV-19/USA/MI-MDHHS-SC20958/2020|EPI_ISL_507747|2</td><td>····················</td><td>····················</td><td>····················</td><td>····················</td><td>NNNNNNNNNNNNNNNNNNNN</td><td>NNNNNNNNNNNNNNNNNNNN</td><td>1</td></tr>
<tr><td>F</td><td>7285</td><td>hCoV-19/USA/MI-MDHHS-SC20959/2020|EPI_ISL_507748|2</td><td>····················</td><td>····················</td><td>····················</td><td>····················</td><td>NNNNNNNNNNNNNNNNNNNN</td><td>NNNNNNNNNNNNNNNNNNNN</td><td>1</td></tr>
<tr><td>F</td><td>7286</td><td>hCoV-19/USA/MI-MDHHS-SC20972/2020|EPI_ISL_507760|2</td><td>····················</td><td>····················</td><td>····················</td><td>····················</td><td>NNNNNNNNNNNNNNNNNNNN</td><td>NNNNNNNNNNNNNNNNNNNN</td><td>1</td></tr>
<tr><td>F</td><td>7287</td><td>hCoV-19/USA/MI-MDHHS-SC20973/2020|EPI_ISL_507761|2</td><td>····················</td><td>····················</td><td>····················</td><td>····················</td><td>NNNNNNNNNNNNNNNNNNNN</td><td>NNNNNNNNNNNNNNNNNNNN</td><td>1</td></tr>
<tr><td>F</td><td>7288</td><td>hCoV-19/USA/MI-MDHHS-SC20975/2020|EPI_ISL_507763|2</td><td>····················</td><td>····················</td><td>····················</td><td>····················</td><td>NNNNNNNNNNNNNNNNNNNN</td><td>NNNNNNNNNNNNNNNNNNNN</td><td>1</td></tr>
<tr><td>F</td><td>7289</td><td>hCoV-19/USA/MI-MDHHS-SC20976/2020|EPI_ISL_507764|2</td><td>····················</td><td>····················</td><td>····················</td><td>····················</td><td>NNNNNNNNNNNNNNNNNNNN</td><td>NNNNNNNNNNNNNNNNNNNN</td><td>1</td></tr>
<tr><td>F</td><td>7344</td><td>hCoV-19/Spain/EX-IBV-005009/2020|EPI_ISL_510318|20</td><td>····················</td><td>····················</td><td>····················</td><td>····················</td><td>NNNNNNNNNNNNNNNNNNNN</td><td>NNNNNNNNNNNNNNNNNNNN</td><td>1</td></tr>
<tr><td>F</td><td>7345</td><td>hCoV-19/Spain/EX-IBV-005010/2020|EPI_ISL_510319|20</td><td>····················</td><td>····················</td><td>····················</td><td>····················</td><td>NNNNNNNNNNNNNNNNNNNN</td><td>NNNNNNNNNNNNNNNNNNNN</td><td>1</td></tr>
<tr><td>F</td><td>7346</td><td>hCoV-19/Spain/EX-IBV-005016/2020|EPI_ISL_510325|20</td><td>····················</td><td>····················</td><td>····················</td><td>····················</td><td>··NNNNN·············</td><td>····················</td><td>1</td></tr>
<tr><td>F</td><td>7347</td><td>hCoV-19/Spain/EX-IBV-005017/2020|EPI_ISL_510326|20</td><td>····················</td><td>····················</td><td>····················</td><td>····················</td><td>NNNNNNNNNNNNNNNNNNNN</td><td>NNNNNNNNNNNNNNNNNNNN</td><td>1</td></tr>
<tr><td>F</td><td>7348</td><td>hCoV-19/Spain/EX-IBV-005018/2020|EPI_ISL_510327|20</td><td>····················</td><td>····················</td><td>····················</td><td>····················</td><td>NNNNNNNNNNNNNNNNNNNN</td><td>NNNNNNNNNNNNNNNNNNNN</td><td>1</td></tr>
<tr><td>F</td><td>7350</td><td>hCoV-19/Spain/EX-IBV-005020/2020|EPI_ISL_510329|20</td><td>····················</td><td>····················</td><td>····················</td><td>····················</td><td>NNNNNNNNNNNNNNNN····</td><td>NNNNNNNNNNNNNNN·····</td><td>1</td></tr>
<tr><td>F</td><td>7351</td><td>hCoV-19/Spain/EX-IBV-005021/2020|EPI_ISL_510330|20</td><td>····················</td><td>····················</td><td>····················</td><td>····················</td><td>NNNNNNNNNNNNNNNNNNNN</td><td>NNNNNNNNNNNNNNNNNNNN</td><td>1</td></tr>
<tr><td>F</td><td>7353</td><td>hCoV-19/Spain/AR-IBV-005025/2020|EPI_ISL_510334|20</td><td>····················</td><td>····················</td><td>····················</td><td>····················</td><td>NNNNNNNNNNNNNNNNNNNN</td><td>NNNNNNNNNNNNNNNNNNNN</td><td>1</td></tr>
<tr><td>F</td><td>7354</td><td>hCoV-19/Spain/AR-IBV-005034/2020|EPI_ISL_510338|20</td><td>····················</td><td>····················</td><td>····················</td><td>····················</td><td>NNNNNNNNNNNNNNNNNNNN</td><td>NNNNNNNNNNNNNNNNNNNN</td><td>1</td></tr>
<tr><td>F</td><td>7355</td><td>hCoV-19/Spain/AR-IBV-005037/2020|EPI_ISL_510340|20</td><td>····················</td><td>····················</td><td>····················</td><td>····················</td><td>NNNNNNNNNNNNNNNNNNNN</td><td>NNNNNNNNNNNNNNNNN···</td><td>1</td></tr>
<tr><td>F</td><td>7356</td><td>hCoV-19/Spain/AR-IBV-005040/2020|EPI_ISL_510343|20</td><td>····················</td><td>····················</td><td>····················</td><td>····················</td><td>NNNNNNNNNNNNNNNNNNNN</td><td>NNNNNNNNNNNNNNNNNNNN</td><td>1</td></tr>
<tr><td>F</td><td>7357</td><td>hCoV-19/Spain/AR-IBV-005041/2020|EPI_ISL_510344|20</td><td>····················</td><td>····················</td><td>····················</td><td>····················</td><td>NNNNNNNNNNNNNNNNNNNN</td><td>NNNNNNNNNNNNNNNNNNNN</td><td>1</td></tr>
<tr><td>F</td><td>7358</td><td>hCoV-19/Spain/AR-IBV-005043/2020|EPI_ISL_510346|20</td><td>····················</td><td>····················</td><td>····················</td><td>····················</td><td>NNNNNNNNNNNNNNNNNNNN</td><td>NNNNNNNNNNNNNNNNNNNN</td><td>1</td></tr>
<tr><td>F</td><td>7359</td><td>hCoV-19/Spain/AR-IBV-005049/2020|EPI_ISL_510349|20</td><td>····················</td><td>····················</td><td>····················</td><td>····················</td><td>NNNNNNNNNNNNNNNNNNNN</td><td>NNNNNNNNNNNNNNNNNNNN</td><td>1</td></tr>
<tr><td>F</td><td>7360</td><td>hCoV-19/Spain/AR-IBV-005051/2020|EPI_ISL_510351|20</td><td>····················</td><td>····················</td><td>····················</td><td>····················</td><td>NNNNNNNNNNNNNNNNNNNN</td><td>NNNNNNNNNNNNNNNNNNNN</td><td>1</td></tr>
<tr><td>F</td><td>7361</td><td>hCoV-19/Spain/AR-IBV-005053/2020|EPI_ISL_510352|20</td><td>····················</td><td>····················</td><td>····················</td><td>····················</td><td>NNNNNNNNNNNNNNNNNNNN</td><td>NNNNNNNNNNNNNNNNNNNN</td><td>1</td></tr>
<tr><td>F</td><td>7362</td><td>hCoV-19/Spain/AR-IBV-005059/2020|EPI_ISL_510354|20</td><td>····················</td><td>····················</td><td>····················</td><td>····················</td><td>NNNNNNNNNNNNNNNNNNNN</td><td>NNNNNNNNNNNNNNNNNNNN</td><td>1</td></tr>
<tr><td>F</td><td>7363</td><td>hCoV-19/Spain/AR-IBV-005062/2020|EPI_ISL_510356|20</td><td>····················</td><td>····················</td><td>····················</td><td>····················</td><td>NNNNNNNNNNNNNNNNNNNN</td><td>NNNNNNNNNNNNNNNNNNNN</td><td>1</td></tr>
<tr><td>F</td><td>7365</td><td>hCoV-19/Spain/AR-IBV-005069/2020|EPI_ISL_510358|20</td><td>····················</td><td>····················</td><td>····················</td><td>····················</td><td>NNNNNNNNNNNNNNNNNNNN</td><td>NNNNNNNNNNNNNNNNNNNN</td><td>1</td></tr>
<tr><td>F</td><td>7366</td><td>hCoV-19/Spain/AR-IBV-005072/2020|EPI_ISL_510359|20</td><td>····················</td><td>····················</td><td>····················</td><td>····················</td><td>NNNNNNNNNNNNNNNNNNNN</td><td>····················</td><td>1</td></tr>
<tr><td>F</td><td>7367</td><td>hCoV-19/Spain/AR-IBV-005074/2020|EPI_ISL_510360|20</td><td>····················</td><td>····················</td><td>····················</td><td>····················</td><td>NNNNNNN·············</td><td>····················</td><td>1</td></tr>
<tr><td>F</td><td>7369</td><td>hCoV-19/Spain/AR-IBV-005082/2020|EPI_ISL_510365|20</td><td>····················</td><td>····················</td><td>····················</td><td>····················</td><td>NNNNNNNNNNNNNNNNNNNN</td><td>NNNNNNNNNNNNNNNNNNNN</td><td>1</td></tr>
<tr><td>F</td><td>7370</td><td>hCoV-19/Spain/AR-IBV-005083/2020|EPI_ISL_510366|20</td><td>····················</td><td>····················</td><td>····················</td><td>····················</td><td>NNNNNNNNNNNNNNNNNNNN</td><td>NNNNNNNNNNNNNNNNNNNN</td><td>1</td></tr>
<tr><td>F</td><td>7371</td><td>hCoV-19/Spain/AR-IBV-005084/2020|EPI_ISL_510367|20</td><td>····················</td><td>····················</td><td>····················</td><td>····················</td><td>NNNNNNNNNNNNNNNNNNNN</td><td>NNNNNNNNNNNNNNNNNNNN</td><td>1</td></tr>
<tr><td>F</td><td>7430</td><td>hCoV-19/South Africa/KRISP-0567/2020|EPI_ISL_49810</td><td>····················</td><td>····················</td><td>····················</td><td>····················</td><td>NNNNNNNNNNNNNNNNNNNN</td><td>NNNNNNNNNNNNNNNNNNNN</td><td>1</td></tr>
<tr><td>F</td><td>7466</td><td>hCoV-19/Wales/PHWC-1692B3/2020|EPI_ISL_499427|2020</td><td>····················</td><td>····················</td><td>····················</td><td>····················</td><td>NNNNNNNNNNNNNNNNN···</td><td>····················</td><td>1</td></tr>
<tr><td>F</td><td>7489</td><td>hCoV-19/Wales/PHWC-1694F9/2020|EPI_ISL_499451|2020</td><td>····················</td><td>····················</td><td>····················</td><td>····················</td><td>NNNNNNNNNNNN········</td><td>····················</td><td>1</td></tr>
<tr><td>F</td><td>7493</td><td>hCoV-19/Wales/PHWC-169550/2020|EPI_ISL_499455|2020</td><td>····················</td><td>····················</td><td>····················</td><td>····················</td><td>·NNNNNNNNNNNNNNNNNNN</td><td>NNNNNNNNNNNNNNNNNNNN</td><td>1</td></tr>
<tr><td>F</td><td>7499</td><td>hCoV-19/England/LIVE-A5902/2020|EPI_ISL_499462|202</td><td>····················</td><td>····················</td><td>····················</td><td>····················</td><td>·······NNNNNNNNNNNNN</td><td>NNNNNNNNNNNNNNNNNNNN</td><td>1</td></tr>
<tr><td>F</td><td>7500</td><td>hCoV-19/England/LIVE-A5C72/2020|EPI_ISL_499463|202</td><td>····················</td><td>····················</td><td>····················</td><td>····················</td><td>·······NNNNNNNNNNNNN</td><td>NNNNNNNNNNNNNNNNNNNN</td><td>1</td></tr>
<tr><td>F</td><td>7501</td><td>hCoV-19/England/LIVE-A5D06/2020|EPI_ISL_499464|202</td><td>····················</td><td>····················</td><td>····················</td><td>····················</td><td>·······NNNNNNNNNNNNN</td><td>NNNNNNNNNNNNNNNNNNNN</td><td>1</td></tr>
<tr><td>F</td><td>7582</td><td>hCoV-19/Spain/EX-IBV-004990/2020|EPI_ISL_510305|20</td><td>····················</td><td>····················</td><td>····················</td><td>····················</td><td>NNNNNNNNNNNNNNNNNNNN</td><td>NNNNNNNNNNNNNNNNNNNN</td><td>1</td></tr>
<tr><td>F</td><td>7583</td><td>hCoV-19/Spain/EX-IBV-004991/2020|EPI_ISL_510306|20</td><td>····················</td><td>····················</td><td>····················</td><td>····················</td><td>NNNNNNNNNNNNNNNNNNNN</td><td>····················</td><td>1</td></tr>
<tr><td>F</td><td>7584</td><td>hCoV-19/Spain/EX-IBV-004992/2020|EPI_ISL_510307|20</td><td>····················</td><td>····················</td><td>····················</td><td>····················</td><td>NNNNNNNNNNNNNNNNNNNN</td><td>NNNNNNNNNNNNNNNNNNNN</td><td>1</td></tr>
<tr><td>F</td><td>7585</td><td>hCoV-19/Spain/EX-IBV-004998/2020|EPI_ISL_510309|20</td><td>····················</td><td>····················</td><td>····················</td><td>····················</td><td>NNNNNNNNNNNNNNNNNNNN</td><td>NNNNNNNNNNNNNNNNNNNN</td><td>1</td></tr>
<tr><td>F</td><td>7586</td><td>hCoV-19/Spain/EX-IBV-005000/2020|EPI_ISL_510310|20</td><td>····················</td><td>····················</td><td>····················</td><td>····················</td><td>NNNNNNNNNNNNNNNNNNNN</td><td>NNNNNNNNNNNNNNNNNNNN</td><td>1</td></tr>
<tr><td>F</td><td>7587</td><td>hCoV-19/Spain/EX-IBV-005001/2020|EPI_ISL_510311|20</td><td>····················</td><td>····················</td><td>····················</td><td>····················</td><td>NNNNNNNNNNNNNNNNNNNN</td><td>NNNNNNNNNNNNNNNNNNNN</td><td>1</td></tr>
<tr><td>F</td><td>7588</td><td>hCoV-19/Spain/EX-IBV-005003/2020|EPI_ISL_510312|20</td><td>····················</td><td>····················</td><td>····················</td><td>····················</td><td>·NNNNNN·············</td><td>····················</td><td>1</td></tr>
<tr><td>F</td><td>7589</td><td>hCoV-19/Spain/EX-IBV-005004/2020|EPI_ISL_510313|20</td><td>····················</td><td>····················</td><td>····················</td><td>····················</td><td>NNNNNNNNNNNNNNNNNNNN</td><td>NNNNN···············</td><td>1</td></tr>
<tr><td>F</td><td>7590</td><td>hCoV-19/Spain/EX-IBV-005005/2020|EPI_ISL_510314|20</td><td>····················</td><td>····················</td><td>····················</td><td>····················</td><td>NNNNNNNNNNNNNNNNNNNN</td><td>NN·····NNNNNNNNNNNNN</td><td>1</td></tr>
<tr><td>F</td><td>7591</td><td>hCoV-19/Spain/EX-IBV-005006/2020|EPI_ISL_510315|20</td><td>····················</td><td>····················</td><td>····················</td><td>····················</td><td>·NNNNNNNNNNNNNNNNNNN</td><td>·············NN·····</td><td>1</td></tr>
<tr><td>F</td><td>7592</td><td>hCoV-19/Spain/EX-IBV-005007/2020|EPI_ISL_510316|20</td><td>····················</td><td>····················</td><td>····················</td><td>····················</td><td>NNNNNNNNNNNNNNNNNNNN</td><td>····················</td><td>1</td></tr>
<tr><td>F</td><td>7595</td><td>hCoV-19/Spain/EX-IBV-005012/2020|EPI_ISL_510321|20</td><td>····················</td><td>····················</td><td>····················</td><td>····················</td><td>NNNNNNNNNNNNNNNNNNNN</td><td>NNNNNNNNNNNNNNNNNNNN</td><td>1</td></tr>
<tr><td>F</td><td>7596</td><td>hCoV-19/Spain/EX-IBV-005013/2020|EPI_ISL_510322|20</td><td>····················</td><td>····················</td><td>····················</td><td>····················</td><td>NNNNNNNNNNNNNNNNNNNN</td><td>NNNNNNNNNNNNNNNNNNNN</td><td>1</td></tr>
<tr><td>F</td><td>7598</td><td>hCoV-19/Spain/EX-IBV-005015/2020|EPI_ISL_510324|20</td><td>····················</td><td>····················</td><td>····················</td><td>····················</td><td>NNNNNNNNNNNNNNNNNNNN</td><td>NNNNNNNNNNNNNNNNNNNN</td><td>1</td></tr>
<tr><td>F</td><td>7745</td><td>hCoV-19/Spain/AR-IBV-005088/2020|EPI_ISL_510371|20</td><td>····················</td><td>····················</td><td>····················</td><td>····················</td><td>NNNNNNNNNNNNNNNNNNNN</td><td>NNNNNNNNNNNNNNN··NNN</td><td>1</td></tr>
<tr><td>F</td><td>7746</td><td>hCoV-19/Spain/AR-IBV-005089/2020|EPI_ISL_510372|20</td><td>····················</td><td>····················</td><td>····················</td><td>····················</td><td>··NN················</td><td>NNNNNNN·············</td><td>1</td></tr>
<tr><td>F</td><td>7747</td><td>hCoV-19/Spain/AR-IBV-005090/2020|EPI_ISL_510373|20</td><td>····················</td><td>····················</td><td>····················</td><td>····················</td><td>NNNNNNNNNNNNNNNNNNNN</td><td>NNNNNNNNNNNNNNNNNNNN</td><td>1</td></tr>
<tr><td>F</td><td>7748</td><td>hCoV-19/Spain/AR-IBV-005091/2020|EPI_ISL_510374|20</td><td>····················</td><td>····················</td><td>····················</td><td>····················</td><td>NNNNNNNNNNNNNNNNNNNN</td><td>NNNNNNNNNNNNNNNNNNNN</td><td>1</td></tr>
<tr><td>F</td><td>7749</td><td>hCoV-19/Spain/AR-IBV-005094/2020|EPI_ISL_510375|20</td><td>····················</td><td>····················</td><td>····················</td><td>····················</td><td>NNNNNNNNNNNNNNNNNNNN</td><td>NNNNNNNNNNNNNNNNNNNN</td><td>1</td></tr>
<tr><td>F</td><td>7750</td><td>hCoV-19/Spain/AR-IBV-005095/2020|EPI_ISL_510376|20</td><td>····················</td><td>····················</td><td>····················</td><td>····················</td><td>·NNNNNN·············</td><td>····················</td><td>1</td></tr>
<tr><td>F</td><td>7751</td><td>hCoV-19/Spain/AR-IBV-005096/2020|EPI_ISL_510377|20</td><td>····················</td><td>····················</td><td>····················</td><td>····················</td><td>·NNNNNNNNNNNNNNNNNNN</td><td>NNN·················</td><td>1</td></tr>
<tr><td>F</td><td>7752</td><td>hCoV-19/Spain/AR-IBV-005097/2020|EPI_ISL_510378|20</td><td>····················</td><td>····················</td><td>····················</td><td>····················</td><td>NNNNNNNNNNNNNNNNNNNN</td><td>NNNNNNNNNNNNNNNNNNNN</td><td>1</td></tr>
<tr><td>F</td><td>7753</td><td>hCoV-19/Spain/AR-IBV-005099/2020|EPI_ISL_510379|20</td><td>····················</td><td>····················</td><td>····················</td><td>····················</td><td>NNNNNNNNNNNNNNNNNNNN</td><td>NNNNNNNNNNNNNNNNNNNN</td><td>1</td></tr>
<tr><td>F</td><td>7755</td><td>hCoV-19/Spain/AR-IBV-005116/2020|EPI_ISL_510383|20</td><td>····················</td><td>····················</td><td>····················</td><td>····················</td><td>NNNNNNNNNNNNNNNNNNNN</td><td>NNNNNNNNNNNNNNN·····</td><td>1</td></tr>
<tr><td>F</td><td>7756</td><td>hCoV-19/Spain/AR-IBV-005119/2020|EPI_ISL_510385|20</td><td>····················</td><td>····················</td><td>····················</td><td>····················</td><td>NNNNNNNNNNNNNNNNNNNN</td><td>NNNNNNNNNNNNNNNNNNNN</td><td>1</td></tr>
<tr><td>F</td><td>7757</td><td>hCoV-19/Spain/AR-IBV-005121/2020|EPI_ISL_510386|20</td><td>····················</td><td>····················</td><td>····················</td><td>····················</td><td>NNNNNNNNNNNNNNNNNNNN</td><td>NNNNNNNNNNNNNNNNNNNN</td><td>1</td></tr>
<tr><td>F</td><td>7758</td><td>hCoV-19/Spain/AR-IBV-005123/2020|EPI_ISL_510387|20</td><td>····················</td><td>····················</td><td>····················</td><td>····················</td><td>NNNNNNNNNNNNNNNNNNNN</td><td>NNNNNNNNNNNNNNN··NNN</td><td>1</td></tr>
<tr><td>F</td><td>7759</td><td>hCoV-19/Spain/AR-IBV-005124/2020|EPI_ISL_510388|20</td><td>····················</td><td>····················</td><td>····················</td><td>····················</td><td>NNNNNNNNNNNNNNNNNNNN</td><td>NNNNNNNNNNNNNNNNNNNN</td><td>1</td></tr>
<tr><td>F</td><td>7761</td><td>hCoV-19/Spain/PV-IBV-004272/2020|EPI_ISL_510391|20</td><td>····················</td><td>····················</td><td>····················</td><td>····················</td><td>NNNNNNN·············</td><td>····················</td><td>1</td></tr>
<tr><td>F</td><td>7764</td><td>hCoV-19/Spain/PV-IBV-004275/2020|EPI_ISL_510394|20</td><td>····················</td><td>····················</td><td>····················</td><td>····················</td><td>NNNNNNNNNNNNNNNNNNNN</td><td>NNNNNNNNNNNNNNNNNNNN</td><td>1</td></tr>
<tr><td>F</td><td>7765</td><td>hCoV-19/Spain/PV-IBV-004277/2020|EPI_ISL_510395|20</td><td>····················</td><td>····················</td><td>····················</td><td>····················</td><td>NNNNNNN·············</td><td>NNNNNNNNNNNNNNN·····</td><td>1</td></tr>
<tr><td>F</td><td>7766</td><td>hCoV-19/Spain/PV-IBV-004278/2020|EPI_ISL_510396|20</td><td>····················</td><td>····················</td><td>····················</td><td>····················</td><td>NNNNNNNNNNNNNNNNNNNN</td><td>NNNNNNNNNNNNNNNNNNNN</td><td>1</td></tr>
<tr><td>F</td><td>7767</td><td>hCoV-19/Spain/PV-IBV-004279/2020|EPI_ISL_510397|20</td><td>····················</td><td>····················</td><td>····················</td><td>····················</td><td>NNNNNNNNNNNNNNNNNNNN</td><td>····················</td><td>1</td></tr>
<tr><td>F</td><td>7768</td><td>hCoV-19/Spain/PV-IBV-004281/2020|EPI_ISL_510398|20</td><td>····················</td><td>····················</td><td>····················</td><td>····················</td><td>NNNNNNNNNNNNNNNNNNNN</td><td>NNNNNNNNNNNNNNNNNNNN</td><td>1</td></tr>
<tr><td>F</td><td>7769</td><td>hCoV-19/Spain/PV-IBV-004282/2020|EPI_ISL_510399|20</td><td>····················</td><td>····················</td><td>····················</td><td>····················</td><td>·NNNNNN·············</td><td>NNNNNNNNNNNNNNN·····</td><td>1</td></tr>
<tr><td>F</td><td>7770</td><td>hCoV-19/Spain/PV-IBV-004283/2020|EPI_ISL_510400|20</td><td>····················</td><td>····················</td><td>····················</td><td>····················</td><td>NNNNNNNNNNNNNNNNNNNN</td><td>NNNNNNNNNNNNNNNNNNNN</td><td>1</td></tr>
</table>

| | | | | | | | | | |
|---|---|---|---|---|---|---|---|---|---|
| F | 7771 | hCoV-19/Spain/PV-IBV-004285/2020\|EPI_ISL_510401\|20 | ···················· | ···················· | ···················· | ···················· | NNNNNNNNNNNNNNNNNNNN | NNNNNNNNNNNNNNNNNNNN | 1 |
| F | 7872 | hCoV-19/USA/WI-UW-1096/2020\|EPI_ISL_516521\|2020-07 | ···················· | ···················· | ···················· | ···················· | NNNNNNNNNNNNNNNNNNNN | NNNNNNNNNNNNNNNNNNNN | 1 |
| F | 7874 | hCoV-19/USA/WI-UW-1098/2020\|EPI_ISL_516523\|2020-07 | ···················· | ···················· | ···················· | ···················· | NNNNNNNNNNNNNNNNNNNN | NNNNNNNNNNNNNNNNNNNN | 1 |
| F | 7895 | hCoV-19/USA/WI-UW-1119/2020\|EPI_ISL_516544\|2020-07 | ···················· | ···················· | ···················· | ···················· | NNNNNNNNNNNNNNNNNNNN | NNNNNNNNNNNNNNNNNNNN | 1 |
| F | 7953 | hCoV-19/Thailand/Bangkok-CONI-0194/2020\|EPI_ISL_49 | ···················· | ···················· | ···················· | ···················· | ··NNNNNNNNNNNNNNNNNN | ···················· | 1 |
| F | 8024 | hCoV-19/South Africa/KRISP-K000586/2020\|EPI_ISL_50 | ···················· | ···················· | ···················· | ···················· | ·NNNNNNNNNNNNNNN···· | ···················· | 1 |
| F | 8041 | hCoV-19/Spain/MD-IBV-005322/2020\|EPI_ISL_510201\|20 | ···················· | ···················· | ···················· | ···················· | NNNNNNNNNNNNNNNNNNNN | NNNNNNNNNNNNNNNNNNNN | 1 |
| F | 8042 | hCoV-19/Spain/MD-IBV-005323/2020\|EPI_ISL_510202\|20 | ···················· | ···················· | ···················· | ···················· | NNNNNNNNNNNNNNNNNNNN | ···················· | 1 |
| F | 8043 | hCoV-19/Spain/MD-IBV-005324/2020\|EPI_ISL_510203\|20 | ···················· | ···················· | ···················· | ···················· | NNNNNNNNNNNNNNNNNNNN | NNNNNNNNNNNNNNN··NNN | 1 |
| F | 8044 | hCoV-19/Spain/MD-IBV-005325/2020\|EPI_ISL_510204\|20 | ···················· | ···················· | ···················· | ···················· | NNNNNNNNNNNNNNNNNNNN | NNNNNNNNNNNNNNNNNNNN | 1 |
| F | 8045 | hCoV-19/Spain/MD-IBV-005326/2020\|EPI_ISL_510205\|20 | ···················· | ···················· | ···················· | ···················· | NNNNNNNNNNNNNNNNNNNN | ···················· | 1 |
| F | 8046 | hCoV-19/Spain/MD-IBV-005328/2020\|EPI_ISL_510207\|20 | ···················· | ···················· | ···················· | ···················· | NNNNNNNNNNNNNNNNNNNN | NNNNNNNNNNNNNNNNNNNN | 1 |
| F | 8047 | hCoV-19/Spain/MD-IBV-005333/2020\|EPI_ISL_510211\|20 | ···················· | ···················· | ···················· | ···················· | NNNNNNNNNNNNNNNNNNNN | NNNNNNNNNNNNNNNNNNNN | 1 |
| F | 8048 | hCoV-19/Spain/MD-IBV-005334/2020\|EPI_ISL_510212\|20 | ···················· | ···················· | ···················· | ···················· | NNNNNNNNNNNNNNNNNNNN | NNNNNNNNNNNNNNNNNNNN | 1 |
| F | 8049 | hCoV-19/Spain/MD-IBV-005335/2020\|EPI_ISL_510213\|20 | ···················· | ···················· | ···················· | ···················· | NNNNNNNNNNNNNNNNNNNN | NNNNNNNNNNNNNNNNNNNN | 1 |
| F | 8050 | hCoV-19/Spain/MD-IBV-005338/2020\|EPI_ISL_510216\|20 | ···················· | ···················· | ···················· | ···················· | NNNNNNNNNNNNNNNNNNNN | ············NNN····· | 1 |
| F | 8051 | hCoV-19/Spain/MD-IBV-005339/2020\|EPI_ISL_510217\|20 | ···················· | ···················· | ···················· | ···················· | NNNNNNNNNNNNNNNNNNNN | NNNNNNNNNNNNNNNNNNNN | 1 |
| F | 8052 | hCoV-19/Spain/MD-IBV-005340/2020\|EPI_ISL_510218\|20 | ···················· | ···················· | ···················· | ···················· | NNNNNNNNNNNNNNNNNNNN | NNNNNNNNNNNNNNNNNNNN | 1 |
| F | 8053 | hCoV-19/Spain/MD-IBV-005343/2020\|EPI_ISL_510220\|20 | ···················· | ···················· | ···················· | ···················· | NNNNNNNNNNNNNNNNNNNN | NNNNNNNNNNNNNNNNNNNN | 1 |
| F | 8054 | hCoV-19/Spain/MD-IBV-005344/2020\|EPI_ISL_510221\|20 | ···················· | ···················· | ···················· | ···················· | NNNNNNNNNNNNNNNNNNNN | NNNNNNNNNNNNNNNNNNNN | 1 |
| F | 8056 | hCoV-19/Spain/MD-IBV-005346/2020\|EPI_ISL_510223\|20 | ···················· | ···················· | ···················· | ···················· | NNNNNNNNNNNNNNNNNNNN | NNNNNNNNNNNNNNNNNNNN | 1 |
| F | 8057 | hCoV-19/Spain/MD-IBV-005347/2020\|EPI_ISL_510224\|20 | ···················· | ···················· | ···················· | ···················· | NNNNNNNNNNNNNNNNNNNN | NNNNNNNNNNNNNNNNNNNN | 1 |
| F | 8058 | hCoV-19/Spain/MD-IBV-005348/2020\|EPI_ISL_510225\|20 | ···················· | ···················· | ···················· | ···················· | NNNNNNNNNNNNNNNNNNNN | NNNNNNNNNNNNNNNNNNNN | 1 |
| F | 8059 | hCoV-19/Spain/MD-IBV-005349/2020\|EPI_ISL_510226\|20 | ···················· | ···················· | ···················· | ···················· | NNNNNNNNNNNNNNNNNNNN | NNNNNNNNNNNNNNNNNNNN | 1 |
| F | 8060 | hCoV-19/Spain/MD-IBV-005350/2020\|EPI_ISL_510227\|20 | ···················· | ···················· | ···················· | ···················· | NNNNNNNNNNNNNNNNNNNN | NNNNNNNNNNNNNNNNNNNN | 1 |
| F | 8061 | hCoV-19/Spain/MD-IBV-005351/2020\|EPI_ISL_510228\|20 | ···················· | ···················· | ···················· | ···················· | NNNNNNNNNNN··NNNNNNN | NNNNNNNNNNNNNNNNN··· | 1 |
| F | 8062 | hCoV-19/Spain/MD-IBV-005354/2020\|EPI_ISL_510231\|20 | ···················· | ···················· | ···················· | ···················· | NNNNNNNNNNNNNNNNNNNN | NNNNNNNNNNNNNNNNNNNN | 1 |
| F | 8063 | hCoV-19/Spain/MD-IBV-005355/2020\|EPI_ISL_510232\|20 | ···················· | ···················· | ···················· | ···················· | NNNNNNNNNNNNNNNNNNNN | NNNNNNNNNNNNNNNNNNNN | 1 |
| F | 8064 | hCoV-19/Spain/MD-IBV-005356/2020\|EPI_ISL_510233\|20 | ···················· | ···················· | ···················· | ···················· | NNNNNNNNNNNNNNNNNNNN | NNNNNNNNNNNNNNNNNNNN | 1 |
| F | 8065 | hCoV-19/Spain/MD-IBV-005360/2020\|EPI_ISL_510236\|20 | ···················· | ···················· | ···················· | ···················· | NNNNNNNNNNNNNNNNNNNN | NNNNNNNNNNNNNNNNNNNN | 1 |
| F | 8066 | hCoV-19/Spain/MD-IBV-005361/2020\|EPI_ISL_510237\|20 | ···················· | ···················· | ···················· | ···················· | NNNNNNNNNNNNNNNNNNNN | NNNNNNNNNNNNNNNNNNNN | 1 |
| F | 8067 | hCoV-19/Spain/MD-IBV-005362/2020\|EPI_ISL_510238\|20 | ···················· | ···················· | ···················· | ···················· | NNNNNNNNNNNNNNNNNNNN | NNNNNNNNNNNNNNNNNNNN | 1 |
| F | 8068 | hCoV-19/Spain/MD-IBV-005365/2020\|EPI_ISL_510240\|20 | ···················· | ···················· | ···················· | ···················· | NNNNNNNNNNNNNNNNNNNN | NNNNNNNNNNNNNNNNNNNN | 1 |
| F | 8069 | hCoV-19/Spain/MD-IBV-005369/2020\|EPI_ISL_510243\|20 | ···················· | ·········A·········· | ···················· | ···················· | NNNNNNNNNNNNNNNNNNNN | NNNNNNNNNNNNNNNNNNNN | 1 |
| F | 8070 | hCoV-19/Spain/MD-IBV-005370/2020\|EPI_ISL_510244\|20 | ···················· | ···················· | ···················· | ···················· | NNNNNNNNNNNNNNNNNNNN | NNNNNNNNNNNNNNNNNNNN | 1 |
| F | 8071 | hCoV-19/Spain/CT-IBV-003150/2020\|EPI_ISL_510247\|20 | ···················· | ···················· | ···················· | ···················· | NNNNNNNNNNNNNNNNNNNN | NNNNNNNNNNNNNNNNNNNN | 1 |
| F | 8142 | hCoV-19/Thailand/Bangkok-CONI-0199/2020\|EPI_ISL_51 | ···················· | ···················· | ···················· | ···················· | NNNNNNNNNNNNNNNNNNNN | ···················· | 1 |
| F | 8143 | hCoV-19/Thailand/Bangkok-CONI-0201/2020\|EPI_ISL_51 | ···················· | ···················· | ···················· | ···················· | ·NNNNNNNNNNN········ | ···················· | 1 |
| F | 8145 | hCoV-19/Thailand/Bangkok-CONI-0206/2020\|EPI_ISL_51 | ···················· | ···················· | ···················· | ···················· | NNNNNNNNNNNNNNNNNNNN | NNNNNNNNNNNNNNNNNNNN | 1 |
| F | 8147 | hCoV-19/Thailand/Bangkok-CONI-0208/2020\|EPI_ISL_51 | ···················· | ···················· | ···················· | ···················· | ··NNNNNNNNNNNNNNNNNN | ···················· | 1 |
| F | 8148 | hCoV-19/Thailand/Bangkok-CONI-0209/2020\|EPI_ISL_51 | ···················· | ···················· | ···················· | ···················· | ··NNNNNNNNNNNNNNNNNN | ···················· | 1 |
| F | 8236 | hCoV-19/Thailand/Bangkok-CONI-0163/2020\|EPI_ISL_49 | ···················· | ·A·················· | ···················· | ···················· | ··NNNNNNNNNNNNNNNNNN | NNNNNNNNNNNNNNNNNNNN | 1 |
| F | 8299 | hCoV-19/Spain/CT-IBV-003152/2020\|EPI_ISL_510249\|20 | ···················· | ···················· | ···················· | ···················· | NNNNNNNNNNNNNNNNNNNN | ···················· | 1 |
| F | 8300 | hCoV-19/Spain/CT-IBV-003153/2020\|EPI_ISL_510250\|20 | ···················· | ···················· | ···················· | ···················· | NNNNNNNNNNNNNNNNNNNN | NNNNNNNNNNNNNNNNNNNN | 1 |
| F | 8301 | hCoV-19/Spain/CT-IBV-003154/2020\|EPI_ISL_510251\|20 | ···················· | ···················· | ···················· | ···················· | ··NNNN·············· | ···················· | 1 |
| F | 8306 | hCoV-19/Spain/CT-IBV-003162/2020\|EPI_ISL_510258\|20 | ···················· | ···················· | ···················· | ···················· | NNNNNNNNNNNNNNNNNNNN | NNNNNNNNNNNNNNNNNNNN | 1 |
| F | 8309 | hCoV-19/Spain/CT-IBV-003165/2020\|EPI_ISL_510261\|20 | ···················· | ···················· | ···················· | ···················· | NNNNNNNNNNNNNNNNNNNN | ·········NNNNNN··NNN | 1 |
| F | 8310 | hCoV-19/Spain/CT-IBV-003166/2020\|EPI_ISL_510262\|20 | ···················· | ···················· | ···················· | ···················· | NNNNNNNNNNNNNNNNNNNN | NNNNNNNNNNNNNNNNNNNN | 1 |
| F | 8312 | hCoV-19/Spain/CT-IBV-003168/2020\|EPI_ISL_510264\|20 | ···················· | ···················· | ···················· | ···················· | NNNNNNNNNNNNNNNNNNNN | NNNNNNNNNNNNNNNNNNNN | 1 |
| F | 8314 | hCoV-19/Spain/CT-IBV-003177/2020\|EPI_ISL_510266\|20 | ···················· | ···················· | ···················· | ···················· | NNNNNNNNNNNNNNNNNNNN | ···················· | 1 |
| F | 8318 | hCoV-19/Spain/GA-IBV-004031/2020\|EPI_ISL_510272\|20 | ···················· | ···················· | ···················· | ···················· | NNNNNNNNNNNNNNNNNNNN | NNNNNNNNNNNNNNNNNNNN | 1 |
| F | 8322 | hCoV-19/Spain/GA-IBV-004038/2020\|EPI_ISL_510277\|20 | ···················· | ···················· | ···················· | ···················· | NNNNNNNNNNNNNNNNNNNN | NNNNN·······NNN····· | 1 |
| F | 8323 | hCoV-19/Spain/MD-IBV-004040/2020\|EPI_ISL_510279\|20 | ···················· | ···················· | ···················· | ···················· | NNNNNNNNNNNNNNNNNNNN | NNNNNNNNNNNNNNNNNNNN | 1 |
| F | 8324 | hCoV-19/Spain/GA-IBV-004041/2020\|EPI_ISL_510280\|20 | ···················· | ···················· | ···················· | ···················· | NNNNNNNNNNN········· | NNNNNNNNNNNNNNN····· | 1 |
| F | 8325 | hCoV-19/Spain/GA-IBV-004042/2020\|EPI_ISL_510281\|20 | ···················· | ···················· | ···················· | ···················· | NNNNNNN············· | ···················· | 1 |
| F | 8328 | hCoV-19/Spain/GA-IBV-004046/2020\|EPI_ISL_510285\|20 | ···················· | ···················· | ···················· | ···················· | ·NNNNNN············· | ···················· | 1 |
| F | 8329 | hCoV-19/Spain/GA-IBV-004047/2020\|EPI_ISL_510286\|20 | ···················· | ···················· | ···················· | ···················· | NNNNNNNNNNNNNNNNNNNN | NNNNNNNNNNNNNNNNNNNN | 1 |
| F | 8330 | hCoV-19/Spain/GA-IBV-004049/2020\|EPI_ISL_510288\|20 | ···················· | ···················· | ···················· | ···················· | NNNNNNNNNNNN········ | ···················· | 1 |
| F | 8333 | hCoV-19/Spain/GA-IBV-004053/2020\|EPI_ISL_510292\|20 | ···················· | ···················· | ···················· | ···················· | NNNNNNN············· | NNNNNNNNNNNNNNN····· | 1 |
| F | 8336 | hCoV-19/Spain/GA-IBV-004058/2020\|EPI_ISL_510296\|20 | ···················· | ···················· | ···················· | ···················· | ·NNNNNN············· | ···················· | 1 |
| F | 8337 | hCoV-19/Spain/GA-IBV-004059/2020\|EPI_ISL_510297\|20 | ···················· | ···················· | ···················· | ···················· | NNNNNNNNNNNNNNNNNNNN | NNNNNNNNNNNNNNNNNNNN | 1 |
| F | 8380 | hCoV-19/SaudiArabia/KAUST-JEDDAH496/2020\|EPI_ISL_5 | ···················· | ···················· | ···················· | ···················· | ·······NNNNNNNNNNNNN | NNNNNNNNNNNNNNNNNNNN | 1 |
| F | 8381 | hCoV-19/SaudiArabia/KAUST-JEDDAH497/2020\|EPI_ISL_5 | ···················· | ···················· | ···················· | ···················· | ···················· | NNNNNNNNNNNNNNNNNNNN | 1 |
| F | 8384 | hCoV-19/Ghana/82879_S48/2020\|EPI_ISL_515100\|2020-0 | ···················· | ···················· | ···················· | ···················· | NNNNNNNNNNNNNNNNNNNN | NNNNNNNNNNNNNNNNNNNN | 1 |
| F | 8385 | hCoV-19/Ghana/83637_S29/2020\|EPI_ISL_515101\|2020-0 | ···················· | ···················· | ···················· | ···················· | NNNNNNNNNNNNNNNNNNNN | NNNNNNNNNNNNNNNN···· | 1 |
| F | 8386 | hCoV-19/Ghana/84049_S31/2020\|EPI_ISL_515102\|2020-0 | ···················· | ···················· | ···················· | ···················· | NNNNNNN············· | NNNNNNN·····NNN····· | 1 |
| F | 8387 | hCoV-19/Ghana/84260_S28/2020\|EPI_ISL_515107\|2020-0 | ···················· | ···················· | ···················· | ···················· | NNNNNNNNNNNNNNNNNNNN | ···················· | 1 |
| F | 8388 | hCoV-19/Ghana/84279_S42/2020\|EPI_ISL_515108\|2020-0 | ···················· | ···················· | ···················· | ···················· | NNNNNNNNNNNNNNNNNNNN | NNNNNNNNNNNNNNN····· | 1 |
| F | 8389 | hCoV-19/Ghana/84430_S51/2020\|EPI_ISL_515110\|2020-0 | ···················· | ···················· | ···················· | ···················· | NNNNNNNNNNNNNNNNNNNN | NNNNNNNNNNNNNNNNNNNN | 1 |
| F | 8390 | hCoV-19/Ghana/84438_S50/2020\|EPI_ISL_515111\|2020-0 | ···················· | ···················· | ···················· | ···················· | NNNNNNNNNNNNNNNNNNNN | ···················· | 1 |
| F | 8429 | hCoV-19/USA/WI-UW-1023/2020\|EPI_ISL_516449\|2020-08 | ···················· | ···················· | ···················· | ···················· | NNNNNNNNNNNNNNNNNNNN | NNNNNNNNNNNNNNNNNNNN | 1 |
| F | 8437 | hCoV-19/USA/WI-UW-1032/2020\|EPI_ISL_516458\|2020-07 | ···················· | ···················· | ···················· | ···················· | NNNNNNNNNNNNNNNNNNNN | NNNNNNNNNNNNNNNNNNNN | 1 |
| F | 8444 | hCoV-19/USA/IL-UW-1039/2020\|EPI_ISL_516465\|2020-08 | ···················· | ···················· | ···················· | ···················· | NNNNNNNNNNNNNNNNNNNN | NNNNNNNNNNNNNNNNNNNN | 1 |
| F | 8501 | hCoV-19/Norway/2857/2020\|EPI_ISL_500780\|2020-07-02 | ···················· | ·A·················· | ···················· | ···················· | NNNNNNNNNN·········· | ···················· | 1 |
| F | 8587 | hCoV-19/Spain/MD-IBV-006363/2020\|EPI_ISL_510106\|20 | ···················· | ···················· | ···················· | ···················· | NNNNNNNNNNNNNNNNNNNN | NNNNNNNNNNNNNNNNNNNN | 1 |
| F | 8588 | hCoV-19/Spain/MD-IBV-006379/2020\|EPI_ISL_510110\|20 | ···················· | ···················· | ···················· | ···················· | NNNNNNNNNNNNNNNNNNNN | NNNNNNNNNNNNNNNNNNNN | 1 |
| F | 8589 | hCoV-19/Spain/MD-IBV-006380/2020\|EPI_ISL_510111\|20 | ···················· | ···················· | ···················· | ···················· | NNNNNNNNNNNNNNNNNNNN | NNNNNNNNNNNNNNNNNNNN | 1 |
| F | 8590 | hCoV-19/Spain/MD-IBV-006382/2020\|EPI_ISL_510112\|20 | ···················· | ···················· | ···················· | ···················· | NNNNNNNNNNNNNNNNNNNN | NNNNNNNNNNNNNNNNNNNN | 1 |
| F | 8591 | hCoV-19/Spain/MD-IBV-006384/2020\|EPI_ISL_510113\|20 | ···················· | ···················· | ···················· | ···················· | NNNNNNNNNNNNNNNNNNNN | NNNNNNNNNNNNNNNNNNNN | 1 |
| F | 8592 | hCoV-19/Spain/MD-IBV-006396/2020\|EPI_ISL_510117\|20 | ···················· | ···················· | ···················· | ···················· | NNNNNNNNNNNNNNNNNNNN | NNNNNNNNNNNNNNNNNNNN | 1 |

| | | | | | | | | | |
|---|---|---|---|---|---|---|---|---|---|
| F | 8593 | hCoV-19/Spain/MD-IBV-006404/2020\|EPI_ISL_510122\|20 | ···················· | ···················· | ···················· | ···················· | NNNNNNNNNNNNNNNNNNNN | NNNNNNNNNNNNNNNNNNNN | 1 |
| F | 8803 | hCoV-19/Spain/MD-IBV-006413/2020\|EPI_ISL_510128\|20 | ···················· | ···················· | ···················· | ···················· | NNNNNNNNNNNNNNNNNNNN | NNNNNNNNNNNNNNNNNNNN | 1 |
| F | 8804 | hCoV-19/Spain/MD-IBV-006414/2020\|EPI_ISL_510129\|20 | ···················· | ···················· | ···················· | ···················· | NNNNNNNNNNNNNNNNNNNN | NNNNNNNNNNNNNNNNNNNN | 1 |
| F | 8805 | hCoV-19/Spain/MD-IBV-006416/2020\|EPI_ISL_510131\|20 | ···················· | ···················· | ···················· | ···················· | NNNNNNNNNNNNNNNNNNNN | NNNNNNNNNNNNNNNNNNNN | 1 |
| F | 8806 | hCoV-19/Spain/MD-IBV-006418/2020\|EPI_ISL_510133\|20 | ···················· | ···················· | ···················· | ···················· | NNNNNNNNNNNNNNNNNNNN | NNNNNNNNNNNNNNNNNNNN | 1 |
| F | 8807 | hCoV-19/Spain/MD-IBV-006420/2020\|EPI_ISL_510135\|20 | ···················· | ···················· | ···················· | ···················· | NNNNNNNNNNNNNNNNNNNN | NNNNNNNNNNNNNNNNNNNN | 1 |
| F | 8808 | hCoV-19/Spain/MD-IBV-006433/2020\|EPI_ISL_510145\|20 | ···················· | ···················· | ···················· | ···················· | NNNNNNNNNNNNNNNNNNNN | NNNNNNNNNNNNNNNNNNNN | 1 |
| F | 8809 | hCoV-19/Spain/MD-IBV-006434/2020\|EPI_ISL_510146\|20 | ···················· | ···················· | ···················· | ···················· | NNNNNNNNNNNNNNNNNNNN | NNNNNNNNNNNNNNNNNNNN | 1 |
| F | 8810 | hCoV-19/Spain/MD-IBV-006439/2020\|EPI_ISL_510149\|20 | ···················· | ···················· | ···················· | ···················· | NNNNNNNNNNNNNNNNNNNN | NNNNNNNNNNNNNNNNNNNN | 1 |
| F | 8811 | hCoV-19/env/USA/UT-CDC-1251/2020\|EPI_ISL_510160\|20 | ···················· | ···················· | ···················· | NNNNNNNN············ | ···················· | ···················· | 1 |
| F | 8812 | hCoV-19/Spain/MD-IBV-005279/2020\|EPI_ISL_510165\|20 | ···················· | ···················· | ···················· | ···················· | NNNNNNNNNNNNNNNNNNNN | NNNNNNNNNNNNNNNNNNNN | 1 |
| F | 8813 | hCoV-19/Spain/MD-IBV-005280/2020\|EPI_ISL_510166\|20 | ···················· | ···················· | ···················· | ···················· | ··NNNNN············· | ···················· | 1 |
| F | 8814 | hCoV-19/Spain/MD-IBV-005281/2020\|EPI_ISL_510167\|20 | ···················· | ···················· | ···················· | ···················· | NNNNNNNNNNNNNNNNNNNN | NNNNNNNNNNNNNNNNNNNN | 1 |
| F | 8815 | hCoV-19/Spain/MD-IBV-005282/2020\|EPI_ISL_510168\|20 | ···················· | ···················· | ···················· | ···················· | NNNNNNNNNNNNNNNNNNNN | NNNNNNNNNNNNNNNNNNNN | 1 |
| F | 8817 | hCoV-19/Spain/MD-IBV-005284/2020\|EPI_ISL_510170\|20 | ···················· | ···················· | ···················· | ···················· | NNNNNNNNNNNNNNNNNNNN | ···················· | 1 |
| F | 8818 | hCoV-19/Spain/MD-IBV-005285/2020\|EPI_ISL_510171\|20 | ···················· | ···················· | ···················· | ···················· | NNNNNNNNNNNNNNNNNNNN | NNNNNNNNNNNNNNNNNNNN | 1 |
| F | 8819 | hCoV-19/Spain/MD-IBV-005286/2020\|EPI_ISL_510172\|20 | ···················· | ···················· | ···················· | ···················· | NNNNNNNNNNNNNNNNNNNN | NNNNNNNNNNNNNNNNNNNN | 1 |
| F | 8820 | hCoV-19/Spain/MD-IBV-005287/2020\|EPI_ISL_510173\|20 | ···················· | ···················· | ···················· | ···················· | NNNNNNNNNNNNNNNNNNNN | NNNNNNNNNNNNNNNNNNNN | 1 |
| F | 8821 | hCoV-19/Spain/MD-IBV-005289/2020\|EPI_ISL_510174\|20 | ···················· | ···················· | ···················· | ···················· | NNNNNNNNNNNNNNNNNNNN | NNNNNNNNNNNNNNNNNNNN | 1 |
| F | 8822 | hCoV-19/Spain/MD-IBV-005291/2020\|EPI_ISL_510176\|20 | ···················· | ···················· | ···················· | ···················· | NNNNNNNNNNNNNNNNNNNN | NNNNNNNNNNNNNNNNNNNN | 1 |
| F | 8823 | hCoV-19/Spain/MD-IBV-005293/2020\|EPI_ISL_510178\|20 | ···················· | ···················· | ···················· | ···················· | NNNNNNNNNNNNNNNNNNNN | NNNNNNNNNNNNNNNNNNNN | 1 |
| F | 8824 | hCoV-19/Spain/MD-IBV-005295/2020\|EPI_ISL_510180\|20 | ···················· | ···················· | ···················· | ···················· | NNNNNNNNNNNNNNNNNNNN | NNNNNNNNNNNNNNNNNNNN | 1 |
| F | 8825 | hCoV-19/Spain/MD-IBV-005298/2020\|EPI_ISL_510182\|20 | ···················· | ···················· | ···················· | ···················· | NNNNNNNNNNNNNNNNNNNN | NNN·········NNNNNNNN | 1 |
| F | 8826 | hCoV-19/Spain/MD-IBV-005299/2020\|EPI_ISL_510183\|20 | ···················· | ···················· | ···················· | ···················· | NNNNNNNNNNNNNNNNNNNN | NNNNNNNNNNNNNNNNNNNN | 1 |
| F | 8827 | hCoV-19/Spain/MD-IBV-005302/2020\|EPI_ISL_510185\|20 | ···················· | ···················· | ···················· | ···················· | ···················· | NNNNN··············· | 1 |
| F | 8828 | hCoV-19/Spain/MD-IBV-005305/2020\|EPI_ISL_510186\|20 | ···················· | ···················· | ···················· | ···················· | NNNNNNNNNNNNNNNNNNNN | NNNNNNNNNNNNNNNNNNNN | 1 |
| F | 8829 | hCoV-19/Spain/MD-IBV-005309/2020\|EPI_ISL_510190\|20 | ···················· | ···················· | ···················· | ···················· | NNNNNNNNNNNNNNNNNNNN | NNNNNNNNNNNNNNNNNNNN | 1 |
| F | 8830 | hCoV-19/Spain/MD-IBV-005311/2020\|EPI_ISL_510191\|20 | ···················· | ···················· | ···················· | ···················· | NNNNNNNNNNNNNNNNNNNN | NNNNNNNNNNNNNNNNNNNN | 1 |
| F | 8831 | hCoV-19/Spain/MD-IBV-005312/2020\|EPI_ISL_510192\|20 | ···················· | ···················· | ···················· | ···················· | NNNNNNNNNNNNNNNNNNNN | NNNNNNNNNNNNNNNNNNNN | 1 |
| F | 8832 | hCoV-19/Spain/MD-IBV-005313/2020\|EPI_ISL_510193\|20 | ···················· | ···················· | ···················· | ···················· | NNNNNNNNNNNNNNNNNNNN | NNNNNNNNNNNNNNNNNNNN | 1 |
| F | 8833 | hCoV-19/Spain/MD-IBV-005314/2020\|EPI_ISL_510194\|20 | ···················· | ···················· | ···················· | ···················· | NNNNNNNNNNNNNNNNNNNN | NNNNNNNNNNNNNNNNNNNN | 1 |
| F | 8834 | hCoV-19/Spain/MD-IBV-005315/2020\|EPI_ISL_510195\|20 | ···················· | ···················· | ···················· | ···················· | NNNNNNNNNNNNNNNNNNNN | NNNNNNNNNNNNNNNNNNNN | 1 |
| F | 8835 | hCoV-19/Spain/MD-IBV-005316/2020\|EPI_ISL_510196\|20 | ···················· | ···················· | ···················· | ···················· | NNNNNNNNNNNNNNNNNNNN | NNNNNNNNNNNNNNNNNNNN | 1 |
| F | 8836 | hCoV-19/Spain/MD-IBV-005320/2020\|EPI_ISL_510199\|20 | ···················· | ···················· | ···················· | ···················· | NNNNNNNNNNNNNNNNNNNN | NNNNNNNNNNNNNNN····· | 1 |
| F | 8881 | hCoV-19/Portugal/PT0941/2020\|EPI_ISL_511491\|2020-0 | ···················· | NNNNNNNNNNNNNN·N·N·· | ···················· | ···················· | ···················· | ···················· | 1 |
| F | 9199 | hCoV-19/Ghana/2573_S27/2020\|EPI_ISL_515082\|2020-03 | ···················· | ···················· | ···················· | ···················· | NNNNNNNNNNNNNNNNNNNN | NNNNNNNNNNNNNNNNNNNN | 1 |
| F | 9200 | hCoV-19/Ghana/2709_S25/2020\|EPI_ISL_515084\|2020-03 | ···················· | ···················· | ···················· | ···················· | NNNNNNN············· | ···················· | 1 |
| F | 9203 | hCoV-19/Ghana/81216_S36/2020\|EPI_ISL_515098\|2020-0 | ···················· | ···················· | ···················· | ···················· | NNNNNNNNNNNNNNNNNNNN | ···················· | 1 |
| F | 9301 | hCoV-19/Wales/PHWC-16905F/2020\|EPI_ISL_499386\|2020 | ···················· | ···················· | ···················· | ···················· | ······NNNNNNNNNNNNNN | ···················· | 1 |
| F | 9304 | hCoV-19/Wales/PHWC-16908C/2020\|EPI_ISL_499389\|2020 | ···················· | ···················· | ···················· | ···················· | NNNNNNNNNNNNNNNNNNNN | NNNNNNNNNNNNNNN····· | 1 |
| F | 9412 | hCoV-19/Spain/CT-IBV-004162/2020\|EPI_ISL_510072\|20 | ···················· | ···················· | ···················· | ···················· | NNNNNNNNNNNNNNNNNNNN | NNNNNNNNNNNNNNNNNNNN | 1 |
| F | 9413 | hCoV-19/Spain/CT-IBV-004166/2020\|EPI_ISL_510075\|20 | ···················· | ···················· | ···················· | ···················· | NNNNNNNNNNNNNNNNNNNN | NNNNNNNNNNNNNNNNNNNN | 1 |
| F | 9414 | hCoV-19/Spain/CT-IBV-004168/2020\|EPI_ISL_510077\|20 | ···················· | ···················· | ···················· | ···················· | NNNNNNNNNNNNNNNNNNNN | NNNNNNNNNNNNNNNNNNNN | 1 |
| F | 9539 | hCoV-19/Spain/AN-IBV-004097/2020\|EPI_ISL_510052\|20 | ···················· | ···················· | ···················· | ···················· | NNNNNNNNNNNNNNNNNNNN | NNNNNNNNNNNNNNNNNNNN | 1 |
| F | 9540 | hCoV-19/Spain/AN-IBV-004098/2020\|EPI_ISL_510053\|20 | ···················· | ···················· | ···················· | ···················· | NNNNNNNNNNNNNNNNNNNN | NNNNNNNNNNNNNNNNNNNN | 1 |
| F | 9541 | hCoV-19/Spain/AN-IBV-004106/2020\|EPI_ISL_510055\|20 | ···················· | ···················· | ···················· | ···················· | NNNNNNNNNNNNNNNNNNNN | NNNNNNNNNNNNNNNNNNNN | 1 |
| F | 9543 | hCoV-19/Spain/AN-IBV-004124/2020\|EPI_ISL_510059\|20 | ···················· | ···················· | ···················· | ···················· | NNNNNNNNNNNNNNNNNNNN | NNNNNNNNNNNNNNNNNNNN | 1 |
| F | 9544 | hCoV-19/Spain/CT-IBV-004154/2020\|EPI_ISL_510066\|20 | ···················· | ···················· | ···················· | ···················· | NNNNNNNNNNNNNNNNNNNN | NNNNNNNNNNNNNNNNNNNN | 1 |
| F | 9545 | hCoV-19/Spain/CT-IBV-004155/2020\|EPI_ISL_510067\|20 | ···················· | ···················· | ···················· | ···················· | NNNNNNNNNNNNNNNNNNNN | NNNNNNNNNNNNNNNNNNNN | 1 |
| F | 9546 | hCoV-19/Spain/CT-IBV-004156/2020\|EPI_ISL_510068\|20 | ···················· | ···················· | ···················· | ···················· | NNNNNNNNNNNNNNNNNNNN | NNNNNNNNNNNNNNNNNNNN | 1 |
| F | 9547 | hCoV-19/Spain/CT-IBV-004157/2020\|EPI_ISL_510069\|20 | ···················· | ···················· | ···················· | ···················· | NNNNNNNNNNNNNNNNNNNN | NNNNNNNNNNNNNNNNNNNN | 1 |
| F | 9611 | hCoV-19/USA/FL-BPHL-0760/2020\|EPI_ISL_512676\|2020- | ···················· | ···················· | ···················· | ···················· | NNNNNNNNNNNNNNNNNNNN | NNNNNNNNNNNNNNNNNNNN | 1 |
| G | 178 | hCoV-19/Australia/VIC7169/2020\|EPI_ISL_521450\|2020 | ···················· | ···················· | ···················· | ···················· | NNNNNNNNNNNNNNNNNNNN | ···················· | 1 |
| G | 208 | hCoV-19/Australia/VIC2639/2020\|EPI_ISL_521485\|2020 | ···················· | ···················· | ···················· | ···················· | NNNNNNNNNNNNNNNNNNNN | ···················· | 1 |
| G | 222 | hCoV-19/Australia/TAS77/2020\|EPI_ISL_522748\|2020-0 | ···················· | ···················· | ···················· | ···················· | NNNNNNNNNNNNNNNNNNNN | ···················· | 1 |
| G | 393 | hCoV-19/England/LIVE-A93E2/2020\|EPI_ISL_526372\|202 | ···················· | ···M················ | ···················· | ···················· | ·······NNNNNNNNNNNNN | NNNNNNNNNNNNNNNNNNNN | 1 |
| G | 404 | hCoV-19/England/LIVE-A9643/2020\|EPI_ISL_526383\|202 | ···················· | ···················· | ···················· | ···················· | ·······NNNNNNNNNNNNN | NNNNNNNNNNNNNNNNNNNN | 1 |
| G | 436 | hCoV-19/USA/MN-MDH-1560/2020\|EPI_ISL_527625\|2020-0 | ···················· | ·A·················· | ···················· | ···················· | ··············NNNNNN | ···················· | 1 |
| G | 623 | hCoV-19/Australia/TAS261/2020\|EPI_ISL_522689\|2020- | ···················· | ···················· | ···················· | ···················· | NNNNNNNNNNNNNNNNNNNN | NNNNNNNNNNNNNNNNNNNN | 1 |
| G | 624 | hCoV-19/Australia/TAS262/2020\|EPI_ISL_522690\|2020- | ···················· | ···················· | ···················· | ···················· | NNNNNNNNNNNNNNNNNNNN | ···················· | 1 |
| G | 626 | hCoV-19/Australia/TAS264/2020\|EPI_ISL_522692\|2020- | ···················· | ···················· | ···················· | ···················· | NNNNNNNNNNNNNNNNNNNN | ···················· | 1 |
| G | 627 | hCoV-19/Australia/TAS265/2020\|EPI_ISL_522693\|2020- | ···················· | ···················· | ···················· | ···················· | NNNN················ | ··············NN···· | 1 |
| G | 628 | hCoV-19/Australia/TAS266/2020\|EPI_ISL_522694\|2020- | ···················· | ···················· | ···················· | ···················· | NNNNNNNNNNNNNNNNNNNN | NNNNNNNNN·NNNNNNNNNN | 1 |
| G | 629 | hCoV-19/Australia/TAS267/2020\|EPI_ISL_522695\|2020- | ···················· | ···················· | ···················· | ···················· | NNNNNNNNNNNNNNNNNNNN | ···················· | 1 |
| G | 632 | hCoV-19/Australia/TAS270/2020\|EPI_ISL_522699\|2020- | ···················· | ···················· | ···················· | ···················· | NNNNNNNNNNNNNNNNNNNN | ···················· | 1 |
| G | 912 | hCoV-19/Australia/TAS18/2020\|EPI_ISL_522630\|2020-0 | ···················· | ···················· | ···················· | ···················· | NNNNNNNNNNNNNNNNNNNN | NNNNNNNNNN······W··· | 1 |
| G | 913 | hCoV-19/Australia/TAS19/2020\|EPI_ISL_522638\|2020-0 | ···················· | ···················· | ···················· | ···················· | NNNNNNNNNNNNNNNNNNNN | NNNNNNNNNNNNNNNNNNNN | 1 |
| G | 914 | hCoV-19/Australia/TAS21/2020\|EPI_ISL_522646\|2020-0 | ···················· | ···················· | ···················· | ···················· | NNNNNNNNNNNNNNNNNNNN | NNNNNNNNNNNNNNNNNNNN | 1 |
| G | 915 | hCoV-19/Australia/TAS220/2020\|EPI_ISL_522650\|2020- | ···················· | ···················· | ···················· | ···················· | NNNNNNNNNNNNNNNNNNNN | NNNNNNNNNNNNNNNNNNNN | 1 |
| G | 916 | hCoV-19/Australia/TAS223/2020\|EPI_ISL_522652\|2020- | ···················· | ···················· | ···················· | ···················· | ···················· | NNNNNNNNNNNNNNNNNNNN | 1 |
| G | 917 | hCoV-19/Australia/TAS224/2020\|EPI_ISL_522653\|2020- | ···················· | ···················· | ···················· | ···················· | NNNNNNNNNNNNNNNNNNNN | ···················· | 1 |
| G | 918 | hCoV-19/Australia/TAS225/2020\|EPI_ISL_522654\|2020- | ···················· | ···················· | ···················· | ···················· | NNNNNNNNNNNNNNNNNNNN | ···················· | 1 |
| G | 919 | hCoV-19/Australia/TAS226/2020\|EPI_ISL_522655\|2020- | ···················· | ···················· | ···················· | ···················· | ···················· | ·······NNNNNNNN····· | 1 |
| G | 920 | hCoV-19/Australia/TAS227/2020\|EPI_ISL_522656\|2020- | ···················· | ···················· | ···················· | ···················· | NNNNNNNNNNNNNNNNNNNN | ···················· | 1 |
| G | 921 | hCoV-19/Australia/TAS228/2020\|EPI_ISL_522657\|2020- | ···················· | ···················· | ···················· | ···················· | NNNNNNNNNNNN········ | ···················· | 1 |
| G | 922 | hCoV-19/Australia/TAS230/2020\|EPI_ISL_522659\|2020- | ···················· | ···················· | ···················· | ···················· | NNNNNNNNNNNN········ | ···················· | 1 |
| G | 923 | hCoV-19/Australia/TAS233/2020\|EPI_ISL_522662\|2020- | ···················· | ···················· | ···················· | ···················· | NNNNNNNNNNNNNNNNNN·· | ···················· | 1 |
| G | 925 | hCoV-19/Australia/TAS237/2020\|EPI_ISL_522666\|2020- | ···················· | ···················· | ···················· | ···················· | NNNNNNNNNNNNNNNNNNNN | N··················· | 1 |
| G | 926 | hCoV-19/Australia/TAS240/2020\|EPI_ISL_522668\|2020- | ···················· | ···················· | ···················· | ···················· | NNNNNNNNNNNNNNNNNNNN | NNNNNNNNNNNNNNNNNNNN | 1 |
| G | 927 | hCoV-19/Australia/TAS241/2020\|EPI_ISL_522669\|2020- | ···················· | ···················· | ···················· | ···················· | NNNNNNNNNNNNNNNNNNNN | NNNNNNNNNNNNNNNNNNNN | 1 |

| | | | | | | | | | |
|---|---|---|---|---|---|---|---|---|---|
| G | 929 | hCoV-19/Australia/TAS245/2020\|EPI_ISL_522672\|2020- | ···················· | ···················· | ···················· | ···················· | NNNNNNNNNNNNNNNNN··· | ···················· | 1 |
| G | 931 | hCoV-19/Australia/TAS247/2020\|EPI_ISL_522674\|2020- | ···················· | ···················· | ···················· | ···················· | NNNNNNNNNNNNNNNNNNNN | NNNNNNNNNNNNNNNNNNNN | 1 |
| G | 932 | hCoV-19/Australia/TAS248/2020\|EPI_ISL_522675\|2020- | ···················· | ···················· | ···················· | ···················· | NNNNNNNNNNN········· | ···················· | 1 |
| G | 933 | hCoV-19/Australia/TAS249/2020\|EPI_ISL_522676\|2020- | ···················· | ···················· | ···················· | ···················· | NNNNNNNNNNNNNNNNNNNN | NNNNNNNNNNNNNNNNNNNN | 1 |
| G | 937 | hCoV-19/Australia/TAS253/2020\|EPI_ISL_522681\|2020- | ···················· | ···················· | ···················· | ···················· | NNNNNNNNNNNNNNNNNNNN | NNNNNNNNNNNNNNNNNNNN | 1 |
| G | 1433 | hCoV-19/Australia/TAS01/2020\|EPI_ISL_522556\|2020-0 | ···················· | ···················· | ···················· | ···················· | NNNNNNNNNNNNNNNNNNNN | NNNNNNNNNNNNNNNNNNNN | 1 |
| G | 1434 | hCoV-19/Australia/TAS03/2020\|EPI_ISL_522558\|2020-0 | ···················· | ···················· | ···················· | ···················· | NNNNNNNNNNNNNNNNNNNN | NNNNNNNNNNNNNNNNNNNN | 1 |
| G | 1435 | hCoV-19/Australia/TAS04/2020\|EPI_ISL_522559\|2020-0 | ···················· | ···················· | ···················· | ···················· | NNNNNNNNNNNNNNNNNNNN | NNNNNNNNNNNNNNNNNNNN | 1 |
| G | 1436 | hCoV-19/Australia/TAS08/2020\|EPI_ISL_522563\|2020-0 | ···················· | ···················· | ···················· | ···················· | NNNNNNNNNNNNNNNNNNNN | NNNNNNNNNNNNNNNNNNNN | 1 |
| G | 1437 | hCoV-19/Australia/TAS09/2020\|EPI_ISL_522564\|2020-0 | ···················· | ···················· | ···················· | ···················· | NNNNNNNNNNNNNNNNNNNN | NNNNNNNNNNNNNNNNNNNN | 1 |
| G | 1438 | hCoV-19/Australia/TAS10/2020\|EPI_ISL_522565\|2020-0 | ···················· | ···················· | ···················· | ···················· | NNNNNNNNNNNNNNNNNNNN | NNNNNNNNNNNNNNNNNNNN | 1 |
| G | 1439 | hCoV-19/Australia/TAS102/2020\|EPI_ISL_522568\|2020- | ···················· | ···················· | ···················· | ···················· | NNNNNNNNNNNNNNNNNNNN | ···················· | 1 |
| G | 1440 | hCoV-19/Australia/TAS109/2020\|EPI_ISL_522574\|2020- | ···················· | ···················· | ···················· | ···················· | NNNNNNNNNNNNNNNNNNNN | NNNNNNNNN··NNNNNN··· | 1 |
| G | 1441 | hCoV-19/Australia/TAS11/2020\|EPI_ISL_522575\|2020-0 | ···················· | ···················· | ···················· | ···················· | NNNNNNNNNNNNNNNNNNNN | NNNNNNNNNNNNNNNNNNNN | 1 |
| G | 1442 | hCoV-19/Australia/TAS110/2020\|EPI_ISL_522576\|2020- | ···················· | ···················· | ···················· | ···················· | NNNNNNNNNNNNNNNNNNNN | NNNNNNNNNNNNNNNNNNNN | 1 |
| G | 1443 | hCoV-19/Australia/TAS133/2020\|EPI_ISL_522599\|2020- | ···················· | ···················· | ···················· | ···················· | NNNNNNNNNNNNNNNNNNNN | NNNNNNNNNNNNNNNNNNNN | 1 |
| G | 1447 | hCoV-19/Australia/TAS275/2020\|EPI_ISL_522704\|2020- | ···················· | ···················· | ···················· | ···················· | NNNNNNNNNNNNNNNNNNNN | ···N················ | 1 |
| G | 1449 | hCoV-19/Australia/TAS37/2020\|EPI_ISL_522714\|2020-0 | ···················· | ···················· | ···················· | ···················· | NNNNNNNNNNNNNNNNNNNN | NNNNNNNNN·········NN | 1 |
| G | 1450 | hCoV-19/Australia/TAS38/2020\|EPI_ISL_522715\|2020-0 | ···················· | ···················· | ···················· | ···················· | NNNNNNNNNNNNNNNNNNNN | NNNNNNNNNNNNNNNNNNNN | 1 |
| G | 1451 | hCoV-19/Australia/TAS39/2020\|EPI_ISL_522716\|2020-0 | ···················· | ···················· | ···················· | ···················· | NNNNNNNNNNNNNNNNNNNN | NNNNNNNNNNNNNNNNNNNN | 1 |
| G | 1452 | hCoV-19/Australia/TAS53/2020\|EPI_ISL_522728\|2020 | ···················· | ···················· | ···················· | ···················· | NNNNNNNNNNNNNNNNNNNN | NNNNNNNNNNNNNNNNNNNN | 1 |
| G | 1453 | hCoV-19/Australia/TAS57/2020\|EPI_ISL_522731\|2020-0 | ···················· | ···················· | ···················· | ···················· | NNNNNNNNNNNNNNNNNNNN | NNNNNNNNNNNNNNNNNNNN | 1 |
| G | 1454 | hCoV-19/Australia/TAS62/2020\|EPI_ISL_522734\|2020-0 | ···················· | ···················· | ···················· | ···················· | NNNNNNNNNNNNNNNNNNNN | NNNNNNNNNNNNNNNNNNNN | 1 |
| G | 1455 | hCoV-19/Australia/TAS65/2020\|EPI_ISL_522737\|2020-0 | ···················· | ···················· | ···················· | ···················· | NNNNNNNNNNNNNNNNNNNN | NNNNNNNNNNNNNNNNNNNN | 1 |
| G | 1456 | hCoV-19/Australia/TAS68/2020\|EPI_ISL_522740\|2020-0 | ···················· | ···················· | ···················· | ···················· | NNNNNNNNNNNNNNNNNNNN | NNNNNNNNNNNNNNNNNNNN | 1 |
| G | 1800 | hCoV-19/USA/FL-BPHL-1024/2020\|EPI_ISL_517914\|2020- | ···················· | ···················· | ···················· | ···················· | ·NNNNNNNNNNNNN······ | ···················· | 1 |
| G | 1813 | hCoV-19/USA/FL-BPHL-1040/2020\|EPI_ISL_517930\|2020- | ···················· | ···················· | ···················· | ···················· | NNNNNNNNNNNNNNNNNNNN | NNNNNNNNNNNN········ | 1 |
| G | 1818 | hCoV-19/USA/FL-BPHL-1046/2020\|EPI_ISL_517936\|2020- | ···················· | ···················· | ···················· | ···················· | NNNNNNNNNNNNNNNNNNNN | NNNNNNNNNNNNNNNNNNNN | 1 |
| G | 1819 | hCoV-19/USA/FL-BPHL-1047/2020\|EPI_ISL_517937\|2020- | ···················· | ···················· | ···················· | ···················· | NNNNNNNNNNNNNNNNNNNN | NNNNNNNNNNNNNNNNNNNN | 1 |
| G | 1826 | hCoV-19/USA/FL-BPHL-1057/2020\|EPI_ISL_517947\|2020- | ···················· | ···················· | ···················· | ···················· | NNNN················ | ···················· | 1 |
| G | 1835 | hCoV-19/Australia/VIC3430/2020\|EPI_ISL_521100\|2020 | ···················· | ···················· | ···················· | ···················· | NNNNNNNNNNNNNNNNNNNN | NNNNNNNNNNNNNNNNNNNN | 1 |
| G | 1836 | hCoV-19/Australia/VIC3431/2020\|EPI_ISL_521101\|2020 | ···················· | ···················· | ···················· | ···················· | ···················· | NNNNNNNNNNNNNNN····· | 1 |
| G | 1840 | hCoV-19/Australia/VIC3440/2020\|EPI_ISL_521106\|2020 | ···················· | ···················· | ···················· | ···················· | NNNN················ | ···················· | 1 |
| G | 1844 | hCoV-19/Australia/VIC3445/2020\|EPI_ISL_521111\|2020 | ···················· | ···················· | ···················· | ···················· | NNNNNNNNNNNNNNNNNNNN | NNNNNNNNNNNNNNNNNNNN | 1 |
| G | 1856 | hCoV-19/Australia/VIC3462/2020\|EPI_ISL_521125\|2020 | ···················· | ···················· | ···················· | ···················· | NNNN················ | ···················· | 1 |
| G | 1860 | hCoV-19/Australia/VIC3473/2020\|EPI_ISL_521134\|2020 | ···················· | ···················· | ···················· | ···················· | ··········GT·NNNNNNN | ···················· | 1 |
| G | 2050 | hCoV-19/USA/FL-BPHL-1012/2020\|EPI_ISL_517902\|2020- | ···················· | ···················· | ···················· | ···················· | NNNNNNNNNNNNNNNNNNNN | NNNNNNNNNNNNNNNNNNNN | 1 |
| G | 2052 | hCoV-19/USA/FL-BPHL-1020/2020\|EPI_ISL_517910\|2020- | ···················· | ···················· | ···················· | ···················· | NNNNNNNNNNNNNNNNNNNN | NNNNNNNNNNNNNN···NNN | 1 |
| G | 2060 | hCoV-19/Australia/VIC3477/2020\|EPI_ISL_521138\|2020 | ···················· | ···················· | ···················· | ···················· | NNNN················ | ···················· | 1 |
| G | 2064 | hCoV-19/Australia/VIC3481/2020\|EPI_ISL_521142\|2020 | ···················· | ···················· | ···················· | ···················· | NNNNNNNNNNNNNNNNNNNN | NNNNNNNNNNNNNNNNNNNN | 1 |
| G | 2067 | hCoV-19/Australia/VIC3484/2020\|EPI_ISL_521145\|2020 | ···················· | ···················· | ···················· | ···················· | NNNNNNNNNNNNNNNNNNNN | ···················· | 1 |
| G | 2068 | hCoV-19/Australia/VIC3485/2020\|EPI_ISL_521146\|2020 | ···················· | ···················· | ···················· | ···················· | NNNNNNNNNNNNNNNNNNNN | NNNNNNNNNNNNNNNNNNNN | 1 |
| G | 2073 | hCoV-19/Australia/VIC3492/2020\|EPI_ISL_521152\|2020 | ···················· | ···················· | ···················· | ···················· | NNNNNNNNNNNNNNNNNNNN | NNNNNNNNNNNNNNNNNNNN | 1 |
| G | 2171 | hCoV-19/England/20168041604/2020\|EPI_ISL_527339\|20 | ···················· | ···················· | ···················· | ···················· | ······NNNNNNNN·N·N·· | ···················· | 1 |
| G | 2403 | hCoV-19/Australia/VIC1991/2020\|EPI_ISL_521897\|2020 | ···················· | ···················· | ···················· | ···················· | NNNNNNNNNNNNNNNNNNNN | NNNNNNNN············ | 1 |
| G | 2404 | hCoV-19/Australia/VIC1992/2020\|EPI_ISL_521898\|2020 | ···················· | ···················· | ···················· | ···················· | ··NNNNNNNNNNNNNNNNNN | NNNNNNNNNNNNNNNNNNNN | 1 |
| G | 2562 | hCoV-19/England/LIVE-A752A/2020\|EPI_ISL_517033\|202 | ···················· | ···················· | ···················· | ···················· | ·······NNNNNNNNNNNNN | ···················· | 1 |
| G | 2563 | hCoV-19/England/LIVE-A7539/2020\|EPI_ISL_517034\|202 | ···················· | ···················· | ···················· | ···················· | ·······NNNNNNNNNNNNN | NNNNNNNNNNNNNNNNNNNN | 1 |
| G | 2564 | hCoV-19/England/LIVE-A7557/2020\|EPI_ISL_517036\|202 | ···················· | ···················· | ···················· | ···················· | ·······NNNNNNNNNNNNN | ···················· | 1 |
| G | 2617 | hCoV-19/Australia/VIC2007/2020\|EPI_ISL_521904\|2020 | ···················· | ···················· | ···················· | ···················· | NNNNNNNNNNNNNNNNNNNN | ···················· | 1 |
| G | 2629 | hCoV-19/Australia/VIC2171/2020\|EPI_ISL_521920\|2020 | ···················· | ···················· | ···················· | ···················· | ···················· | NNNNNNNNN··········· | 1 |
| G | 2819 | hCoV-19/England/LIVE-A7302/2020\|EPI_ISL_517002\|202 | ···················· | ···················· | ···················· | ···················· | ·······NNNNNNNNNNNNN | NNNNNNNNNNNNNNNNNNNN | 1 |
| G | 2820 | hCoV-19/England/LIVE-A7311/2020\|EPI_ISL_517003\|202 | ···················· | ···················· | ···················· | ···················· | ·······NNNNNNNNNNNNN | NNNNNNNNNNNNNNNNNNNN | 1 |
| G | 2822 | hCoV-19/England/LIVE-A733F/2020\|EPI_ISL_517005\|202 | ···················· | ···················· | ···················· | ···················· | ·······NNNNNNNNNNNNN | ···················· | 1 |
| G | 2823 | hCoV-19/England/LIVE-A734E/2020\|EPI_ISL_517006\|202 | ···················· | ···················· | ···················· | ···················· | ·······NNNNNNNNNNNNN | NNNNNNNNNNNNNNNNNNNN | 1 |
| G | 2824 | hCoV-19/England/LIVE-A735D/2020\|EPI_ISL_517007\|202 | ···················· | ···················· | ···················· | ···················· | ·······NNNNNNNNNNNNN | ···················· | 1 |
| G | 2826 | hCoV-19/England/LIVE-A738A/2020\|EPI_ISL_517010\|202 | ···················· | ···················A | ···················· | ···················· | ·······NNNNNNNNNNNNN | NNNNNNNNNNNNNNNNNNNN | 1 |
| G | 2829 | hCoV-19/England/LIVE-A73B7/2020\|EPI_ISL_517013\|202 | ···················· | ···················· | ···················· | ···················· | ·······NNNNNNNNNNNNN | NNNNNNNNNNNNNNNNNNNN | 1 |
| G | 2830 | hCoV-19/England/LIVE-A73C6/2020\|EPI_ISL_517014\|202 | ···················· | ···················· | ···················· | ···················· | ·······NNNNNNNNNNNNN | NNNNNNNNNNNNNNNNNNNN | 1 |
| G | 2832 | hCoV-19/England/LIVE-A73E4/2020\|EPI_ISL_517016\|202 | ···················· | ···················· | ···················· | ···················· | ·······NNNNNNNNNNNNN | NNNNNNNNNNNNNNNNNNNN | 1 |
| G | 2833 | hCoV-19/England/LIVE-A73F3/2020\|EPI_ISL_517017\|202 | ···················· | ···················· | ···················· | ···················· | ·······NNNNNNNNNNNNN | NNNNNNNNNNNNNNNNNNNN | 1 |
| G | 2834 | hCoV-19/England/LIVE-A740F/2020\|EPI_ISL_517018\|202 | ···················· | ···················· | ···················· | ···················· | ·······NNNNNNNNNNNNN | ···················· | 1 |
| G | 2837 | hCoV-19/England/LIVE-A744B/2020\|EPI_ISL_517022\|202 | ···················· | ···················· | ···················· | ···················· | ·······NNNNNNNNNNNNN | ···················· | 1 |
| G | 2838 | hCoV-19/England/LIVE-A745A/2020\|EPI_ISL_517023\|202 | ···················· | ···················· | ···················· | ···················· | ·······NNNNNNNNNNNNN | ···················· | 1 |
| G | 2841 | hCoV-19/England/LIVE-A74A5/2020\|EPI_ISL_517027\|202 | ···················· | ···················· | ···················· | ···················· | ·······NNNNNNNNNNNNN | ···················· | 1 |
| G | 2842 | hCoV-19/England/LIVE-A74C3/2020\|EPI_ISL_517029\|202 | ···················· | ···················· | ···················· | ···················· | ·······NNNNNNNNNNNNN | ···················· | 1 |
| G | 2843 | hCoV-19/England/LIVE-A7584/2020\|EPI_ISL_517039\|202 | ···················· | ···················· | ···················· | ···················· | ·······NNNNNNNNNNNNN | NNNNNNNNNNNNNNNNNNNN | 1 |
| G | 2979 | hCoV-19/Australia/VIC3923/2020\|EPI_ISL_520635\|2020 | ···················· | ···················· | ···················· | ···················· | NNNNNNNNNNNNNNNNNNNN | ···················· | 1 |
| G | 2988 | hCoV-19/Australia/VIC3933/2020\|EPI_ISL_520645\|2020 | ···················· | ···················· | ···················· | ···················· | NNNNNNNNNNNNNNNNNNNN | ···················· | 1 |
| G | 2993 | hCoV-19/Australia/VIC3939/2020\|EPI_ISL_520650\|2020 | ···················· | ···················· | ···················· | ···················· | NNNN················ | ···················· | 1 |
| G | 2997 | hCoV-19/Australia/VIC3943/2020\|EPI_ISL_520654\|2020 | ···················· | ···················· | ···················· | ···················· | NNNN················ | ···················· | 1 |
| G | 3002 | hCoV-19/Australia/VIC3949/2020\|EPI_ISL_520660\|2020 | ···················· | ···················· | ···················· | ···················· | NNNNNNNNNNNNNNNNNNNN | ···················· | 1 |
| G | 3042 | hCoV-19/Australia/VIC2235/2020\|EPI_ISL_521972\|2020 | ···················· | ···················· | ···················· | ···················· | NNNNNNNNNNNNNNNNNNNN | NNNNNNNNNNNNNNNNNNNN | 1 |
| G | 3043 | hCoV-19/Australia/VIC2236/2020\|EPI_ISL_521973\|2020 | ···················· | ···················· | ···················· | ···················· | NNNNNNNNNNNNNNNNNNNN | NNNNNNNNNNNNNNNNNNNN | 1 |
| G | 3044 | hCoV-19/Australia/VIC2237/2020\|EPI_ISL_521974\|2020 | ···················· | ···················· | ···················· | ···················· | NNNNNNNNNNNNNNNNNNNN | NNNNNNNNNNNNNNNNNNNN | 1 |
| G | 3045 | hCoV-19/Australia/VIC2242/2020\|EPI_ISL_521975\|2020 | ···················· | ···················· | ···················· | ···················· | NNNNNNNNNNNNNNNNNNNN | NNNNNNNNNNNNNNNNNNNN | 1 |
| G | 3046 | hCoV-19/Australia/VIC2243/2020\|EPI_ISL_521976\|2020 | ···················· | ···················· | ···················· | ···················· | ···················· | NNNNNNNNNNNNNNN····· | 1 |
| G | 3047 | hCoV-19/Australia/VIC2244/2020\|EPI_ISL_521977\|2020 | ···················· | ···················· | ···················· | ···················· | NNNNNNNNNNNNNNNNNNNN | NNNNNNNNNNNNNNNNNNNN | 1 |
| G | 3049 | hCoV-19/Australia/VIC2250/2020\|EPI_ISL_521981\|2020 | ···················· | ···················· | ···················· | ···················· | NNNNNNNNNNNNNNNNNNNN | ···················· | 1 |
| G | 3057 | hCoV-19/Australia/VIC2270/2020\|EPI_ISL_521992\|2020 | ···················· | ···················· | ···················· | ···················· | NNNNNNNNNNNNNNNNNNNN | ·············NNNNNNN | 1 |

| | | | | | | | | | |
|---|---|---|---|---|---|---|---|---|---|
| G | 3060 | hCoV-19/Australia/VIC2273/2020\|EPI_ISL_521995\|2020 | ···················· | ···················· | ···················· | ···················· | NNNN················ | ···················· | 1 |
| G | 3098 | hCoV-19/Australia/VIC6145/2020\|EPI_ISL_518156\|2020 | ···················· | ···················· | ···················· | ···················· | NNNNNNNNNNNNNNNNNNNN | ···················· | 1 |
| G | 3158 | hCoV-19/Australia/VIC3895/2020\|EPI_ISL_520607\|2020 | ···················· | ···················· | ···················· | ···················· | NNNNNNNNNNNNNNNNNNNN | NNNNNNNNNNNNNNNNNNNN | 1 |
| G | 3163 | hCoV-19/Australia/VIC2948/2020\|EPI_ISL_521778\|2020 | ···················· | ···················· | ···················· | ···················· | NNNN················ | ···················· | 1 |
| G | 3173 | hCoV-19/Australia/VIC2958/2020\|EPI_ISL_521788\|2020 | ···················· | ···················· | ···················· | ···················· | ·NNNNNNNNNNNNNNNNNNN | ···················· | 1 |
| G | 3184 | hCoV-19/Australia/VIC2184/2020\|EPI_ISL_521930\|2020 | ···················· | ···················· | ···················· | ···················· | NNNNNNNNNNNNNNNNNNNN | NNNNNNNNNNNNNNNNNNNN | 1 |
| G | 3186 | hCoV-19/Australia/VIC2186/2020\|EPI_ISL_521932\|2020 | ···················· | ···················· | ···················· | ···················· | NNNNNNNNNNNNNNNNNNNN | NNNNNNNNNNNNNNNNNNNN | 1 |
| G | 3238 | hCoV-19/USA/FL-BPHL-1126/2020\|EPI_ISL_526612\|2020- | ···················· | ···················· | ···················· | ···················· | ···················· | NNNNN······NNNN·NNNN | 1 |
| G | 3259 | hCoV-19/USA/FL-BPHL-1148/2020\|EPI_ISL_526634\|2020- | ···················· | ···················· | ···················· | ···················· | NNNNNNNNNNNNNNNNNNNN | ············NNNNNNNN | 1 |
| G | 3261 | hCoV-19/USA/FL-BPHL-1150/2020\|EPI_ISL_526636\|2020- | ···················· | ···················· | ···················· | ···················· | NNNNNN·············· | ···················· | 1 |
| G | 3268 | hCoV-19/USA/FL-BPHL-1162/2020\|EPI_ISL_526648\|2020- | ···················· | ···················· | ···················· | ···················· | NNNNNNNNNNNNNNNNNNNN | NNNNNN···········NNN | 1 |
| G | 3270 | hCoV-19/USA/FL-BPHL-1165/2020\|EPI_ISL_526651\|2020- | ···················· | ···················· | ···················· | ···················· | NNNNNNNNNNNN········ | NNNNNNNNNNN········· | 1 |
| G | 3273 | hCoV-19/USA/FL-BPHL-1170/2020\|EPI_ISL_526656\|2020- | ···················· | ···················· | ···················· | ···················· | NNNNNNNNNNNNNNNNNNNN | NNNNNNNNNNNNNNNNNNNN | 1 |
| G | 3274 | hCoV-19/USA/FL-BPHL-1173/2020\|EPI_ISL_526659\|2020- | ···················· | ···················· | ···················· | ···················· | NNNNNNNNNNNNNNNN···· | ···················· | 1 |
| G | 3295 | hCoV-19/Switzerland/BL-UHB-42176198/2020\|EPI_ISL_5 | ···················· | ···················· | ···················· | ···················· | NNNNNNNNNNNNNNNNNNNN | NNNNNNNNNNNNNNNNNNNN | 1 |
| G | 3298 | hCoV-19/Switzerland/BS-UHB-42177236/2020\|EPI_ISL_5 | ···················· | ···················· | ···················· | ···················· | ·NNNNNNNNNNNNNNNNNNN | ···················· | 1 |
| G | 3303 | hCoV-19/Switzerland/BS-UHB-42178886/2020\|EPI_ISL_5 | ···················· | ···················· | ···················· | ···················· | NNNNNNNNNNNNNNNNNNNN | NNNNNNNNN··········· | 1 |
| G | 3304 | hCoV-19/Switzerland/BL-UHB-42180261/2020\|EPI_ISL_5 | ···················· | ···················· | ···················· | ···················· | NNNNNNNNNNNNNNNNNNNN | NNNNNNNNNNNNNNNNNNNN | 1 |
| G | 3305 | hCoV-19/Switzerland/BS-UHB-42181090/2020\|EPI_ISL_5 | ···················· | ···················· | ···················· | ···················· | NNNNNNNNNNNNNNNNNNNN | NNNNNNNNNNNNNNNNNNNN | 1 |
| G | 3308 | hCoV-19/Switzerland/BS-UHB-42181850/2020\|EPI_ISL_5 | ···················· | ···················· | ···················· | ···················· | NNNNNNNNNNNNNNNNNNNN | NNNNNNNNNNNNNNNNNNNN | 1 |
| G | 3309 | hCoV-19/Switzerland/BL-UHB-42181880/2020\|EPI_ISL_5 | ···················· | ···················· | ···················· | ···················· | NNNNNNNNNNNNNNNNNNNN | NNNNNNNNNNNNNNNNNNNN | 1 |
| G | 3310 | hCoV-19/Switzerland/BS-UHB-42182611/2020\|EPI_ISL_5 | ···················· | ···················· | ···················· | ···················· | NNNNNNNNNNNNNNNNNNNN | NNNNNNNNNNNNNNNNNNNN | 1 |
| G | 3311 | hCoV-19/Switzerland/BS-UHB-42183075/2020\|EPI_ISL_5 | ···················· | ···················· | ···················· | ···················· | NNNNNNNNNNNNNNNNNNNN | NNNNNNNNNNNNNNNNNNNN | 1 |
| G | 3312 | hCoV-19/Switzerland/BS-UHB-42183137/2020\|EPI_ISL_5 | ···················· | ···················· | ···················· | ···················· | NNNNNNNNNNNNNNNNNNNN | NNNNN··NNN·······NNN | 1 |
| G | 3313 | hCoV-19/Switzerland/BS-UHB-42183147/2020\|EPI_ISL_5 | ···················· | ···················· | ···················· | ···················· | NNNNNNNNNNNNNNNNNNNN | ···················· | 1 |
| G | 3314 | hCoV-19/Switzerland/BS-UHB-42183615/2020\|EPI_ISL_5 | ···················· | ···················· | ···················· | ···················· | NNNNNNNNNNNNNNNNNNNN | NNNNNNNNNNNNNNNNNNNN | 1 |
| G | 3315 | hCoV-19/Switzerland/BS-UHB-42183764/2020\|EPI_ISL_5 | ···················· | ···················· | ···················· | ···················· | NNNNNNNNNNNNNNNNNNNN | NNNNNNNNNNNNNNNNNNNN | 1 |
| G | 3316 | hCoV-19/Switzerland/BL-UHB-42184023/2020\|EPI_ISL_5 | ···················· | ···················· | ···················· | ···················· | NNNNNNNNNNNNNNNNNNNN | NNNNNNNNNN·········· | 1 |
| G | 3317 | hCoV-19/Switzerland/BS-UHB-42184169/2020\|EPI_ISL_5 | ···················· | ···················· | ···················· | ···················· | NNNNNNNNNNNNNNNNNNNN | NNNNNNNNNNNNNNNNNNNN | 1 |
| G | 3319 | hCoV-19/Switzerland/BS-UHB-42184956/2020\|EPI_ISL_5 | ···················· | ···················· | ···················· | ···················· | NNNNNNNNNNNNNNNNNNNN | NNNNNNNNNNNNNNNNNNNN | 1 |
| G | 3321 | hCoV-19/Switzerland/BS-UHB-42185476/2020\|EPI_ISL_5 | ···················· | ···················· | ···················· | ···················· | NNNNNNNNNNNNNNNNNNNN | NNNNNNNNNNNNNNNNNNNN | 1 |
| G | 3322 | hCoV-19/Switzerland/BS-UHB-42185657/2020\|EPI_ISL_5 | ···················· | ···················· | ···················· | ···················· | NNNNNNNNNNNNNNNNNNNN | NNNNNNNNNNNNNNNNNNNN | 1 |
| G | 3323 | hCoV-19/Switzerland/BL-UHB-42185829/2020\|EPI_ISL_5 | ···················· | ···················· | ···················· | ···················· | NNNNNNNNNNNNNNNNNNNN | NNNNNNNNNNNNNNNNNNNN | 1 |
| G | 3324 | hCoV-19/Switzerland/BS-UHB-42185836/2020\|EPI_ISL_5 | ···················· | ···················· | ···················· | ···················· | NNNNNNNNNNNNNNNNNNNN | ···················· | 1 |
| G | 3325 | hCoV-19/Switzerland/BS-UHB-42185864/2020\|EPI_ISL_5 | ···················· | ···················· | ···················· | ···················· | NNNNNNNNNNNNNNNNNNNN | NNNNNNNNNNNNNNNNNNNN | 1 |
| G | 3326 | hCoV-19/Switzerland/BL-UHB-42186158/2020\|EPI_ISL_5 | ···················· | ···················· | ···················· | ···················· | NNNNNNNNNNNNNNNNNNNN | NNNNNNNNNNNNNNNNNNNN | 1 |
| G | 3327 | hCoV-19/Switzerland/BS-UHB-42186169/2020\|EPI_ISL_5 | ···················· | ···················· | ···················· | ···················· | NNNNNNNNNNNNNNNNNNNN | NNNNNNNNNNNNNNNNNNNN | 1 |
| G | 3328 | hCoV-19/Switzerland/BS-UHB-42186205/2020\|EPI_ISL_5 | ···················· | ···················· | ···················· | ···················· | NNNNNNNNNNNNNNNNNNNN | NNNNNNNNNNNNNNNNNNNN | 1 |
| G | 3418 | hCoV-19/Australia/VIC3838/2020\|EPI_ISL_520551\|2020 | ···················· | ···················· | ···················· | ···················· | NNNNNNNNNNN········· | ···················· | 1 |
| G | 3447 | hCoV-19/Australia/NT52/2020\|EPI_ISL_521861\|2020-07 | ···················· | ···················· | ···················· | ···················· | NNNNNNNNNNNNNNNNNNNN | NNNNNNNNN··········· | 1 |
| G | 3551 | hCoV-19/Ireland/D-NVRL-71IRL96201/2020\|EPI_ISL_525 | ···················· | ···················· | ···················· | ···················· | NNNNNNNNNNNNNNNNNNNN | NNNNNNNNNNNNNNNNNNNN | 1 |
| G | 3573 | hCoV-19/USA/FL-BPHL-1190/2020\|EPI_ISL_526676\|2020- | ···················· | ···················· | ···················· | ···················· | NNNNNNNNNNNNNNNNNNNN | NNNNNNNNNNNNNNNNNNNN | 1 |
| G | 3669 | hCoV-19/Australia/VIC3998/2020\|EPI_ISL_519387\|2020 | ···················· | ···················· | ···················· | ···················· | NNNN················ | ···················· | 1 |
| G | 3670 | hCoV-19/Australia/VIC4000/2020\|EPI_ISL_519389\|2020 | ···················· | ···················· | ···················· | ···················· | NNNNNNNNNNNNNNNNNNNN | ···················· | 1 |
| G | 3671 | hCoV-19/Australia/VIC4001/2020\|EPI_ISL_519390\|2020 | ···················· | ···················· | ···················· | ···················· | NNNN················ | ···················· | 1 |
| G | 3714 | hCoV-19/Australia/VIC3596/2020\|EPI_ISL_520342\|2020 | ···················· | ···················· | ···················· | ···················· | NNNNNNNNNNNNNNNNN··· | ···················· | 1 |
| G | 3716 | hCoV-19/Australia/VIC3599/2020\|EPI_ISL_520345\|2020 | ···················· | ···················· | ···················· | ···················· | NNNNNNNNNNNNNNNNNNNN | NNNNNNNNNNNNNNN····· | 1 |
| G | 3717 | hCoV-19/Australia/VIC3601/2020\|EPI_ISL_520346\|2020 | ···················· | ···················· | ···················· | ···················· | NNNN················ | ···················· | 1 |
| G | 3720 | hCoV-19/Australia/VIC3606/2020\|EPI_ISL_520350\|2020 | ···················· | ···················· | ···················· | ···················· | NNNNNNNNNNNNNNNNNNNN | NNNNNNNNNNNNNNNNNNNN | 1 |
| G | 3722 | hCoV-19/Australia/VIC3608/2020\|EPI_ISL_520352\|2020 | ···················· | ···················· | ···················· | ···················· | NNNNNNNNNNNNNNNNNNNN | ················NNNN | 1 |
| G | 3723 | hCoV-19/Australia/VIC3609/2020\|EPI_ISL_520353\|2020 | ···················· | ···················· | ···················· | ···················· | NNNNNNNNNNNNNNNNNNNN | NNNNNNNNNNNNNNNN···· | 1 |
| G | 3725 | hCoV-19/Australia/VIC3613/2020\|EPI_ISL_520356\|2020 | ···················· | ···················· | ···················· | ···················· | NNNNNNNNNNNNNNNNNNNN | ···················· | 1 |
| G | 3738 | hCoV-19/Australia/VIC3629/2020\|EPI_ISL_520370\|2020 | ···················· | ···················· | ···················· | ···················· | NNNN················ | ···················· | 1 |
| G | 3740 | hCoV-19/Australia/VIC3632/2020\|EPI_ISL_520372\|2020 | ···················· | ···················· | ···················· | ···················· | NNNNNNNNNNNNNNNNNNNN | NNNNNNNNNNNNNNNNNNNN | 1 |
| G | 3741 | hCoV-19/Australia/VIC3633/2020\|EPI_ISL_520373\|2020 | ···················· | ···················· | ···················· | ···················· | NNNN················ | ···················· | 1 |
| G | 3744 | hCoV-19/Australia/VIC3637/2020\|EPI_ISL_520377\|2020 | ···················· | ···················· | ···················· | ···················· | NNNN················ | ···················· | 1 |
| G | 3748 | hCoV-19/Australia/VIC3643/2020\|EPI_ISL_520383\|2020 | ···················· | ···················· | ···················· | ···················· | NNNN················ | ···················· | 1 |
| G | 3834 | hCoV-19/Australia/VIC3023/2020\|EPI_ISL_521852\|2020 | ···················· | ···················· | ···················· | ···················· | NNN················· | NNNNNNNNNNNNNNNNNNNN | 1 |
| G | 3873 | hCoV-19/Australia/VIC3576/2020\|EPI_ISL_520325\|2020 | ···················· | ···················· | ···················· | ···················· | NNNN················ | ···················· | 1 |
| G | 3879 | hCoV-19/Australia/VIC3583/2020\|EPI_ISL_520331\|2020 | ···················· | ···················· | ···················· | ···················· | ·············NNNNNNN | ···················· | 1 |
| G | 3886 | hCoV-19/Australia/VIC3592/2020\|EPI_ISL_520338\|2020 | ···················· | ···················· | ···················· | ···················· | NNNN················ | ···················· | 1 |
| G | 3888 | hCoV-19/Australia/VIC3597/2020\|EPI_ISL_520343\|2020 | ···················· | ···················· | ···················· | ···················· | NNNN················ | ···················· | 1 |
| G | 3912 | hCoV-19/Mexico/CMX-INMEGEN-21/2020\|EPI_ISL_522986\| | ···················· | ···················· | ···················· | ···················· | NNNNNNNNNNNNNNNNNNNN | NNNNNNNNNNNNNNNNNNNN | 1 |
| G | 4005 | hCoV-19/Scotland/EDB7185/2020\|EPI_ISL_526521\|2020- | ···················· | ···················· | ···················· | ···················· | NNNNNNNNNNNNNNNNNNNN | NNNNNNNNNNNNNNNNNNNN | 1 |
| G | 4006 | hCoV-19/Scotland/EDB7275/2020\|EPI_ISL_526526\|2020- | ···················· | ···················· | ···················· | ···················· | NNNNNNNNNNNNNNNNNNNN | NNNNNNNNNNNNNNNNNNNN | 1 |
| G | 4010 | hCoV-19/Scotland/EDB7365/2020\|EPI_ISL_526536\|2020- | ···················· | ·A·················· | ···················· | ···················· | NNNNNNNNNNNNNNNNNNNN | NNNNNNNNNNNNNNNNNNNN | 1 |
| G | 4021 | hCoV-19/USA/FL-BPHL-1081/2020\|EPI_ISL_526567\|2020- | ···················· | ···················· | ···················· | ···················· | NNNNNNNNNNNNNNNNNNNN | ···················· | 1 |
| G | 4024 | hCoV-19/USA/FL-BPHL-1084/2020\|EPI_ISL_526570\|2020- | ···················· | ···················· | ···················· | ···················· | NNNNNNNNNNNNNNNNNNNN | ···················· | 1 |
| G | 4030 | hCoV-19/USA/FL-BPHL-1095/2020\|EPI_ISL_526581\|2020- | ···················· | ···················· | ···················· | ···················· | ········NNNNNNNNNNNN | ···················· | 1 |
| G | 4034 | hCoV-19/USA/FL-BPHL-1102/2020\|EPI_ISL_526588\|2020- | ···················· | ···················· | ···················· | ···················· | NNNNNNT············· | ···················· | 1 |
| G | 4046 | hCoV-19/USA/WI-UW-1191/2020\|EPI_ISL_527801\|2020-08 | ···················· | ···················· | ···················· | ···················· | NNNNNNNNNNNNNNNNNNNN | NNNNNNNNNNNNNNNNNNNN | 1 |
| G | 4047 | hCoV-19/USA/WI-UW-1192/2020\|EPI_ISL_527802\|2020-08 | ···················· | ···················· | ···················· | ···················· | NNNNNNNNNNNNNNNNNNNN | NNNNNNNNNNNNNNNNNNNN | 1 |
| G | 4048 | hCoV-19/USA/WI-UW-1193/2020\|EPI_ISL_527803\|2020-08 | ···················· | ···················· | ···················· | ···················· | NNNNNNNNNNNNNNNNNNNN | NNNNNNNNNNNNNNNNNNNN | 1 |
| G | 4049 | hCoV-19/USA/WI-UW-1194/2020\|EPI_ISL_527804\|2020-08 | ···················· | ···················· | ···················· | ···················· | NNNNNNNNNNNNNNNNNNNN | NNNNNNNNNNNNNNNNNNNN | 1 |
| G | 4050 | hCoV-19/USA/WI-UW-1196/2020\|EPI_ISL_527806\|2020-08 | ···················· | ···················· | ···················· | ···················· | NNNNNNNNNNNNNNNNNNNN | NNNNNNNNNNNNNNNNNNNN | 1 |
| G | 4051 | hCoV-19/USA/WI-UW-1198/2020\|EPI_ISL_527808\|2020-08 | ···················· | ···················· | ···················· | ···················· | NNNNNNNNNNNNNNNNNNNN | NNNNNNNNNNNNNNNNNNNN | 1 |
| G | 4076 | hCoV-19/Brazil/SP-226/2020\|EPI_ISL_527868\|2020-04- | ···················· | ···················· | ···················· | ·········NNNNNN····· | ···················· | ···················· | 1 |
| G | 4210 | hCoV-19/Australia/VIC3493/2020\|EPI_ISL_520249\|2020 | ···················· | ···················· | ···················· | ···················· | NNNNNNNNNNNNNNNNNNNN | NNNNNNNNNNNNNNNNNNNN | 1 |
| G | 4217 | hCoV-19/Australia/VIC3501/2020\|EPI_ISL_520257\|2020 | ···················· | ···················· | ···················· | ···················· | NNNN················ | ···················· | 1 |
| G | 4231 | hCoV-19/Australia/VIC3527/2020\|EPI_ISL_520276\|2020 | ···················· | ···················· | ···················· | ···················· | NNNN················ | ···················· | 1 |

| | | | | | | | | |
|---|---|---|---|---|---|---|---|---|
| G | 4238 hCoV-19/Australia/VIC3534/2020\|EPI_ISL_520283\|2020 | ···················· | ···················· | ···················· | ···················· | NNNNNNNNNNNNNNNNNNNN | ···················· | 1 |
| G | 4240 hCoV-19/Australia/VIC3538/2020\|EPI_ISL_520287\|2020 | ···················· | ···················· | ···················· | ···················· | NNNNNNNNNNNNNNNNNNNN | ···················· | 1 |
| G | 4269 hCoV-19/Australia/VIC2752/2020\|EPI_ISL_521587\|2020 | ···················· | ···················· | ···················· | ···················· | NNNN················ | ···················· | 1 |
| G | 4278 hCoV-19/Australia/VIC2762/2020\|EPI_ISL_521597\|2020 | ···················· | ···················· | ···················· | ···················· | ···················· | NNNN···NNNNNNNNNNNNN | 1 |
| G | 4279 hCoV-19/Australia/VIC2764/2020\|EPI_ISL_521599\|2020 | ···················· | ···················· | ···················· | ···················· | ···················· | NNNNNNNNNNNNNNN····· | 1 |
| G | 4336 hCoV-19/Australia/VIC2927/2020\|EPI_ISL_521758\|2020 | ···················· | ···················· | ···················· | ···················· | NNNNNNNNNNNNNNNNNNNN | NNNNNNNNNNNNNNNNNNNN | 1 |
| G | 4342 hCoV-19/Australia/VIC2934/2020\|EPI_ISL_521765\|2020 | ···················· | ···················· | ···················· | ···················· | ·············NNNNNNN | ···················· | 1 |
| G | 4348 hCoV-19/USA/WI-UW-1136/2020\|EPI_ISL_522893\|2020-08 | ···················· | ···················· | ···················· | ···················· | NNNNNNNNNNNNNNNNNNNN | NNNNNNNNNNNNNNNNNNNN | 1 |
| G | 4409 hCoV-19/Australia/VIC3543/2020\|EPI_ISL_520292\|2020 | ···················· | ···················· | ···················· | ···················· | NNNNNNNNNNNNNNNNN··· | ···················· | 1 |
| G | 4443 hCoV-19/Mexico/CMX-INMEGEN-03/2020\|EPI_ISL_522874\| | ···················· | ···················· | ···················· | ···················· | NNNNNNNNNNNNNNNNNNNN | NNNNNNNNNNNNNNNNNNNN | 1 |
| G | 4444 hCoV-19/Mexico/CMX-INMEGEN-06/2020\|EPI_ISL_522877\| | ···················· | ·A·················· | ···················· | ···················· | NNNNNNNNNNNNNNNNNNNN | NNNNNNNNNNNNNNNNNNNN | 1 |
| G | 4445 hCoV-19/Mexico/CMX-INMEGEN-07/2020\|EPI_ISL_522878\| | ···················· | ···················· | ···················· | ···················· | NNNNNNNNNNNNNNNNNNNN | NNNNNNNNNNNNNNNNNNNN | 1 |
| G | 4446 hCoV-19/Mexico/CMX-INMEGEN-09/2020\|EPI_ISL_522880\| | ···················· | ·A·················· | ···················· | ···················· | NNNNNNNNNNNNNNNNNNNN | NNNNNNNNNNNNNNNNNNNN | 1 |
| G | 4450 hCoV-19/USA/WI-UW-1126/2020\|EPI_ISL_522884\|2020-08 | ···················· | ···················· | ···················· | ···················· | NNNNNNNNNNNNNNNNNNNN | NNNNNNNNNNNNNNNNNNNN | 1 |
| G | 4454 hCoV-19/USA/WI-UW-1131/2020\|EPI_ISL_522888\|2020-08 | ···················· | ···················· | ···················· | ···················· | NNNNNNNNNNNNNNNNNNNN | NNNNNNNNNNNNNNNNNNNN | 1 |
| G | 4455 hCoV-19/USA/WI-UW-1132/2020\|EPI_ISL_522889\|2020-08 | ···················· | ···················· | ···················· | ···················· | NNNNNNNNNNNNNNNNNNNN | NNNNNNNNNNNNNNNNNNNN | 1 |
| G | 4456 hCoV-19/Mexico/CMX-INMEGEN-16/2020\|EPI_ISL_522981\| | ···················· | ···················· | ···················· | ···················· | NNNNNNNNNNNNNNNNNNNN | NNNNNNNNNNNNNNNNNNNN | 1 |
| G | 4512 hCoV-19/England/PORT-2D1FCD/2020\|EPI_ISL_526436\|20 | ···················· | ···················· | ···················· | ···················· | NNNNNNNNNNNNNNNNNNNN | NNNNNNNNNNNNNNNNNNNN | 1 |
| G | 4515 hCoV-19/Scotland/EDB1496/2020\|EPI_ISL_526460\|2020- | ···················· | ·A·················· | ···················· | ···················· | NNNNNNNNNNNNNNNNNNNN | NNNNNNNNNNNNNNNNNNNN | 1 |
| G | 4518 hCoV-19/Scotland/EDB1762/2020\|EPI_ISL_526467\|2020- | ···················· | ·A·················· | ···················· | ···················· | NNNNNNNNNNNNNNNNNNNN | NNNNNNNNNNNNNNNNNNNN | 1 |
| G | 4604 hCoV-19/USA/IL-UW-1190/2020\|EPI_ISL_527794\|2020-08 | ···················· | ·N·················· | ···················· | ···················· | NNNNNNNNNNNNNNNNNNNN | NNNNNNNNNNNNNNNNNNNN | 1 |
| G | 4605 hCoV-19/USA/WI-UW-1184/2020\|EPI_ISL_527795\|2020-08 | ···················· | ···················· | ···················· | ···················· | NNNNNNNNNNNNNNNNNNNN | NNNNNNNNNNNNNNNNNNNN | 1 |
| G | 4606 hCoV-19/USA/WI-UW-1185/2020\|EPI_ISL_527796\|2020-08 | ···················· | ···················· | ···················· | ···················· | NNNNNNNNNNNNNNNNNNNN | NNNNNNNNNNNNNNNNNNNN | 1 |
| G | 4607 hCoV-19/USA/WI-UW-1186/2020\|EPI_ISL_527797\|2020-08 | ···················· | ···················· | ···················· | ···················· | NNNNNNNNNNNNNNNNNNNN | NNNNNNNNNNNNNNNNNNNN | 1 |
| G | 4608 hCoV-19/USA/WI-UW-1188/2020\|EPI_ISL_527799\|2020-08 | ···················· | ···················· | ···················· | ···················· | NNNNNNNNNNNNNNNNNNNN | NNNNNNNNNNNNNNNNNNNN | 1 |
| G | 4670 hCoV-19/Australia/VIC3574/2020\|EPI_ISL_520323\|2020 | ···················· | ···················· | ···················· | ···················· | NNNNNNNNNNNNNNNNNNNN | NNNNNNNNNNNNNNNNNNNN | 1 |
| G | 4672 hCoV-19/Australia/VIC2766/2020\|EPI_ISL_521601\|2020 | ···················· | ···················· | ···················· | ···················· | NNNN················ | NNNNNNNNNNNNNNNNNNNN | 1 |
| G | 4680 hCoV-19/Australia/VIC2776/2020\|EPI_ISL_521610\|2020 | ···················· | ···················· | ···················· | ···················· | ··········NNNNNNNNNN | ···················· | 1 |
| G | 4684 hCoV-19/Australia/VIC2780/2020\|EPI_ISL_521614\|2020 | ···················· | ···················· | ···················· | ···················· | ···NNNNNNNNNNNNNNNNN | NNNNNNNNNNNNNNNNNNNN | 1 |
| G | 4686 hCoV-19/Australia/VIC2782/2020\|EPI_ISL_521616\|2020 | ···················· | ···················· | ···················· | ···················· | NNNN················ | ···················· | 1 |
| G | 4687 hCoV-19/Australia/VIC2783/2020\|EPI_ISL_521617\|2020 | ···················· | ···················· | ···················· | ···················· | NNNNNNNNNNNNNNNNNNNN | ···················· | 1 |
| G | 4689 hCoV-19/Australia/VIC2785/2020\|EPI_ISL_521619\|2020 | ···················· | ···················· | ···················· | ···················· | NNNNNNNNNNNNNNNNNNNN | NNNNNNNNNNNNNNNNNNNN | 1 |
| G | 4692 hCoV-19/Australia/VIC2792/2020\|EPI_ISL_521625\|2020 | ···················· | ···················· | ···················· | ···················· | NNNNNNNNNNNNNNNNNNNN | NNNNNNNNNNNNNNNNNNNN | 1 |
| G | 4694 hCoV-19/Australia/VIC2794/2020\|EPI_ISL_521627\|2020 | ···················· | ···················· | ···················· | ···················· | NNNNNNNNNNNNNNNNNNNN | NNNNNNNNNNNNNNNNNNNN | 1 |
| G | 4695 hCoV-19/Australia/VIC2795/2020\|EPI_ISL_521628\|2020 | ···················· | ···················· | ···················· | ···················· | NNNNNNNNNNNNNNNNNNNN | NNNNNNNNNNNNNNNNNNNN | 1 |
| G | 4696 hCoV-19/Australia/VIC2796/2020\|EPI_ISL_521629\|2020 | ···················· | ···················· | ···················· | ···················· | NNNNNNNNNNNNNNNNNNNN | NNNN···NNNNNNNNNN··· | 1 |
| G | 4700 hCoV-19/Australia/VIC2802/2020\|EPI_ISL_521633\|2020 | ···················· | ···················· | ···················· | ···················· | ··NNNNNNNNNNNNNNNNNN | ··············NNNNNN | 1 |
| G | 4701 hCoV-19/Australia/VIC2803/2020\|EPI_ISL_521634\|2020 | ···················· | ···················· | ···················· | ···················· | NNNNNNNNNNNNNNNNNNNN | NNNNNNNNNNNNNNNNNNNN | 1 |
| G | 4709 hCoV-19/Australia/VIC2811/2020\|EPI_ISL_521642\|2020 | ···················· | ···················· | ···················· | ···················· | ···················· | NNNN················ | 1 |
| G | 4716 hCoV-19/Australia/VIC2818/2020\|EPI_ISL_521649\|2020 | ···················· | ···················· | ···················· | ···················· | NNNNNNNNNNNNNN······ | ···················· | 1 |
| G | 4722 hCoV-19/USA/WI-UW-1143/2020\|EPI_ISL_522900\|2020-08 | ···················· | ···················· | ···················· | ···················· | NNNNNNNNNNNNNNNNNNNN | NNNNNNNNNNNNNNNNNNNN | 1 |
| G | 4723 hCoV-19/USA/WI-UW-1144/2020\|EPI_ISL_522901\|2020-08 | ···················· | ···················· | ···················· | ···················· | NNNNNNNNNNNNNNNNNNNN | NNNNNNNNNNNNNNNNNNNN | 1 |
| G | 4724 hCoV-19/USA/WI-UW-1145/2020\|EPI_ISL_522902\|2020-08 | ···················· | ···················· | ···················· | ···················· | NNNNNNNNNNNNNNNNNNNN | NNNNNNNNNNNNNNNNNNNN | 1 |
| G | 4726 hCoV-19/USA/WI-UW-1148/2020\|EPI_ISL_522904\|2020-08 | ···················· | ···················· | ···················· | ···················· | NNNNNNNNNNNNNNNNNNNN | NNNNNNNNNNNNNNNNNNNN | 1 |
| G | 4727 hCoV-19/USA/WI-UW-1149/2020\|EPI_ISL_522905\|2020-08 | ···················· | ···················· | ···················· | ···················· | NNNNNNNNNNNNNNNNNNNN | NNNNNNNNNNNNNNNNNNNN | 1 |
| G | 4728 hCoV-19/USA/WI-UW-1150/2020\|EPI_ISL_522906\|2020-08 | ···················· | ···················· | ···················· | ···················· | NNNNNNNNNNNNNNNNNNNN | NNNNNNNNNNNNNNNNNNNN | 1 |
| G | 4729 hCoV-19/USA/WI-UW-1151/2020\|EPI_ISL_522907\|2020-08 | ···················· | ···················· | ···················· | ···················· | NNNNNNNNNNNNNNNNNNNN | ···················· | 1 |
| G | 4730 hCoV-19/USA/WI-UW-1152/2020\|EPI_ISL_522908\|2020-08 | ···················· | ···················· | ···················· | ···················· | NNNNNNNNNNNNNNNNNNNN | NNNNNNNNNNNNNNNNNNNN | 1 |
| G | 4732 hCoV-19/USA/WI-UW-1154/2020\|EPI_ISL_522910\|2020-08 | ···················· | ·A·················· | ···················· | ···················· | NNNNNNNNNNNNNNNNNNNN | NNNNNNNNNNNNNNNNNNNN | 1 |
| G | 4733 hCoV-19/USA/WI-UW-1155/2020\|EPI_ISL_522911\|2020-08 | ···················· | ···················· | ···················· | ···················· | NNNNNNNNNNNNNNNNNNNN | NNNNNNNNNNNNNNNNNNNN | 1 |
| G | 4734 hCoV-19/USA/WI-UW-1156/2020\|EPI_ISL_522912\|2020-08 | ···················· | ···················· | ···················· | ···················· | NNNNNNNNNNNNNNNNNNNN | NNNNNNNNNNNNNNNNNNNN | 1 |
| G | 4737 hCoV-19/USA/WI-UW-1159/2020\|EPI_ISL_522915\|2020-08 | ···················· | ···················· | ···················· | ···················· | NNNNNNNNNNNNNNNNNNNN | NNNNNNNNNNNNNNNNNNNN | 1 |
| G | 4738 hCoV-19/USA/WI-UW-1161/2020\|EPI_ISL_522917\|2020-08 | ···················· | ···················· | ···················· | ···················· | NNNNNNNNNNNNNNNNNNNN | NNNNNNNNNNNNNNNNNNNN | 1 |
| G | 4739 hCoV-19/USA/WI-UW-1162/2020\|EPI_ISL_522918\|2020-08 | ···················· | ···A················ | ···················· | ···················· | NNNNNNNNNNNNNNNNNNNN | NNNNNNNNNNNNNNNNNNNN | 1 |
| G | 4740 hCoV-19/USA/WI-UW-1168/2020\|EPI_ISL_522924\|2020-08 | ···················· | ···················· | ···················· | ···················· | NNNNNNNNNNNNNNNNNNNN | NNNNNNNNNNNNNNNNNNNN | 1 |
| G | 4741 hCoV-19/USA/WI-UW-1169/2020\|EPI_ISL_522925\|2020-08 | ···················· | ···················· | ···················· | ···················· | NNNNNNNNNNNNNNNNNNNN | NNNNNNNNNNNNNNNNNNNN | 1 |
| G | 4742 hCoV-19/USA/WI-UW-1170/2020\|EPI_ISL_522926\|2020-08 | ···················· | ···················· | ···················· | ···················· | NNNNNNNNNNNNNNNNNNNN | NNNNNNNNNNNNNNNNNNNN | 1 |
| G | 4744 hCoV-19/USA/WI-UW-1173/2020\|EPI_ISL_522928\|2020-08 | ···················· | ···················· | ···················· | ···················· | NNNNNNNNNNNNNNNNNNNN | NNNNNNNNNNNNNNNNNNNN | 1 |
| G | 4746 hCoV-19/USA/WI-UW-1176/2020\|EPI_ISL_522931\|2020-08 | ···················· | ···················· | ···················· | ···················· | NNNNNNNNNNNNNNNNNNNN | NNNNNNNNNNNNNNNNNNNN | 1 |
| G | 4754 hCoV-19/Mexico/CMX-INMEGEN-10/2020\|EPI_ISL_522940\| | ···················· | ·A·················· | ···················· | ···················· | NNNNNNNNNNNNNNNNNNNN | NNNNNNNNNNNNNNNNNNNN | 1 |
| G | 4755 hCoV-19/Mexico/CMX-INMEGEN-11/2020\|EPI_ISL_522941\| | ···················· | ···A················ | ···················· | ···················· | NNNNNNNNNNNNNNNNNNNN | NNNNNNNNNNNNNNNNNNNN | 1 |
| G | 4756 hCoV-19/Mexico/CMX-INMEGEN-12/2020\|EPI_ISL_522942\| | ···················· | ···················· | ···················· | ···················· | NNNNNNNNNNNNNNNNNNNN | NNNNNNNNNNNNNNNNNNNN | 1 |
| G | 4763 hCoV-19/Mexico/CMX-INMEGEN-13/2020\|EPI_ISL_522978\| | ···················· | ···················· | ···················· | ···················· | NNNNNNNNNNNNNNNNNNNN | NNNNNNNNNNNNNNNNNNNN | 1 |
| G | 4764 hCoV-19/Mexico/CMX-INMEGEN-15/2020\|EPI_ISL_522980\| | ···················· | ···················· | ···················· | ···················· | NNNNNNNNNNNNNNNNNNNN | NNNNNNNNNNNNNNNNNNNN | 1 |
| G | 4765 hCoV-19/Mexico/CMX-INMEGEN-17/2020\|EPI_ISL_522982\| | ···················· | ·A·················· | ···················· | ···················· | NNNNNNNNNNNNNNNNNNNN | NNNNNNNNNNNNNNNNNNNN | 1 |
| G | 4766 hCoV-19/Mexico/CMX-INMEGEN-19/2020\|EPI_ISL_522984\| | ···················· | ···················· | ···················· | ···················· | NNNNNNNNNNNNNNNNNNNN | NNNNNNNNNNNNNNNNNNNN | 1 |
| G | 4867 hCoV-19/England/LIVE-AA220/2020\|EPI_ISL_517422\|202 | ···················· | ···················· | ···················· | ···················· | ···················· | NNNNNNNNNNNNNNNNNNNN | 1 |
| G | 4868 hCoV-19/England/LIVE-AA2D5/2020\|EPI_ISL_517432\|202 | ···················· | ···················· | ···················· | ···················· | ·······NNNNNNNNNNNNN | NNNNNNNNNNNNNNNNNNNN | 1 |
| G | 4869 hCoV-19/England/LIVE-AA2E4/2020\|EPI_ISL_517433\|202 | ···················· | ···················· | ···················· | ···················· | ·······NNNNNNNNN···· | ···················· | 1 |
| G | 4889 hCoV-19/Australia/VIC5935/2020\|EPI_ISL_518714\|2020 | ···················· | ···················· | ···················· | ···················· | ···················· | NNNNNNNNNNNNNNNNNNNN | 1 |
| G | 5122 hCoV-19/England/LIVE-A925E/2020\|EPI_ISL_517282\|202 | ···················· | ···················· | ···················· | ···················· | ·······NNNNNNNNNNNNN | NNNNNNNNNNNNNNNNNNNN | 1 |
| G | 5123 hCoV-19/England/LIVE-A927C/2020\|EPI_ISL_517284\|202 | ···················· | ···················· | ···················· | ···················· | ·······NNNNNNNNNNNNN | ···················· | 1 |
| G | 5128 hCoV-19/England/LIVE-A92E5/2020\|EPI_ISL_517291\|202 | ···················· | ···················· | ···················· | ···················· | ·······NNNNNNNNNNNNN | ···················· | 1 |
| G | 5138 hCoV-19/England/LIVE-AA527/2020\|EPI_ISL_517464\|202 | ···················· | ···················· | ···················· | ···················· | ·······NNNNNNNNNNNNN | NNNNNNNNNNNNNNNNNNNN | 1 |
| G | 5139 hCoV-19/England/LIVE-AA536/2020\|EPI_ISL_517465\|202 | ···················· | ···················· | ···················· | ···················· | ·······NNNNNNNNNNNNN | ···················· | 1 |
| G | 5140 hCoV-19/England/LIVE-AA545/2020\|EPI_ISL_517466\|202 | ···················· | ···················· | ···················· | ···················· | ·······NNNNNNNNNNNNN | NNNNNNNNNNNNNNNNNNNN | 1 |
| G | 5149 hCoV-19/England/LIVE-AA642/2020\|EPI_ISL_517479\|202 | ···················· | ···················· | ···················· | ···················· | ·······NNNNNNNNNNNN· | ···················· | 1 |
| G | 5151 hCoV-19/England/LIVE-AA660/2020\|EPI_ISL_517481\|202 | ···················· | ···················· | ···················· | ···················· | ·······NNNNNNNNNNNNN | ···················· | 1 |
| G | 5163 hCoV-19/England/LIVE-AA74F/2020\|EPI_ISL_517495\|202 | ···················· | ···················· | ···················· | ···················· | ·······NNNNNNNNNNNNN | ···················· | 1 |
| G | 5377 hCoV-19/England/LIVE-A9995/2020\|EPI_ISL_517300\|202 | ···················· | ···················· | ···················· | ···················· | ·······NNNNNNNNNNNNN | ···················· | 1 |

| | | | | | | | | |
|---|---|---|---|---|---|---|---|---|
| G | 5381 hCoV-19/England/LIVE-A99D1/2020\|EPI_ISL_517304\|202 | ···················· | ···················· | ···················· | ···················· | ·······NNNNNNNNNNNNN | NNNNNNNNNNNNNNNNNNNN | 1 |
| G | 5383 hCoV-19/England/LIVE-A99FF/2020\|EPI_ISL_517306\|202 | ···················· | ···················· | ···················· | ···················· | ·······NNNNNNNNNNNNN | ···················· | 1 |
| G | 5393 hCoV-19/England/LIVE-A9A92/2020\|EPI_ISL_517316\|202 | ···················· | ···················· | ···················· | ···················· | ·······NNNNNNNNNNNNN | NNNNNNNNNNNNNNNNNNNN | 1 |
| G | 5394 hCoV-19/England/LIVE-A9AA1/2020\|EPI_ISL_517317\|202 | ···················· | ···················· | ···················· | ···················· | ·······NNNNNNNNNNN·· | NNNNNNNNNNNNNNNNNNNN | 1 |
| G | 5442 hCoV-19/Australia/VIC5852/2020\|EPI_ISL_518645\|2020 | ···················· | ···················· | ···················· | ···················· | NNNNNNNNNNNN········ | ···················· | 1 |
| G | 5526 hCoV-19/Australia/VIC3218/2020\|EPI_ISL_520903\|2020 | ···················· | ···················· | ···················· | ···················· | NNNNNNNNNNNNNNNNNNNN | ···················· | 1 |
| G | 5556 hCoV-19/Australia/VIC3254/2020\|EPI_ISL_520934\|2020 | ···················· | ···················· | ···················· | ···················· | NNNNNNNNNNNNNNNNNNNN | ···················· | 1 |
| G | 5559 hCoV-19/Australia/VIC3257/2020\|EPI_ISL_520937\|2020 | ···················· | ···················· | ···················· | ···················· | ···················· | NNNNNNNNNNNNNNNNNNNN | 1 |
| G | 5564 hCoV-19/Australia/VIC3263/2020\|EPI_ISL_520942\|2020 | ···················· | ···················· | ···················· | ···················· | NNNNNNNNNNNNNNNNNNNN | NNNNNNNNNNNNNNNNNNNN | 1 |
| G | 5565 hCoV-19/Australia/VIC3264/2020\|EPI_ISL_520943\|2020 | ···················· | ···················· | ···················· | ···················· | NNNNNNNNNNNNNNNNNNNN | NNNNNNNNNNNN········ | 1 |
| G | 5639 hCoV-19/England/LIVE-A8671/2020\|EPI_ISL_517196\|202 | ···················· | ···················· | ···················· | ···················· | ·······NNNNNNNNNNNNN | NNNNNNNNNNNNNNNNNNNN | 1 |
| G | 5640 hCoV-19/England/LIVE-A86AE/2020\|EPI_ISL_517198\|202 | ···················· | ···················· | ···················· | ···················· | ·······NNNNNNNNNNNNN | NNNNNNNNNNNNNNNNNNNN | 1 |
| G | 5641 hCoV-19/England/LIVE-A86BD/2020\|EPI_ISL_517199\|202 | ···················· | ···················· | ···················· | ···················· | ·······NNNNNNNNNNNNN | NNNNNNNNNNNNNNNNNNNN | 1 |
| G | 5646 hCoV-19/England/LIVE-A9B26/2020\|EPI_ISL_517325\|202 | ···················· | ···················A | ···················· | ···················· | ·······NNNNNNNNNNNNN | ···················· | 1 |
| G | 5653 hCoV-19/England/LIVE-A9BAE/2020\|EPI_ISL_517332\|202 | ···················· | ···················· | ···················· | ···················· | ·······NNNNNNNNNNNNN | NNNNNNNNNNNNNNNNNNNN | 1 |
| G | 5655 hCoV-19/England/LIVE-A9BDB/2020\|EPI_ISL_517334\|202 | ···················· | ···················· | ···················· | ···················· | ·······NNNNNNNNNNNNN | ···················· | 1 |
| G | 5661 hCoV-19/England/LIVE-A9C32/2020\|EPI_ISL_517340\|202 | ···················· | ···················· | ···················· | ···················· | ·······NNNNNNNNNNNNN | NNNNNNNNNNNNNNNNNNNN | 1 |
| G | 5663 hCoV-19/England/LIVE-A9C6F/2020\|EPI_ISL_517343\|202 | ···················· | ···················· | ···················· | ···················· | ·······NNNNNNNNNNNNN | ···················· | 1 |
| G | 5675 hCoV-19/England/LIVE-A9D20/2020\|EPI_ISL_517355\|202 | ···················· | ···················· | ···················· | ···················· | ·······NNNNNNNNNNNNN | NNNNNNNNNNNNNNNNNNNN | 1 |
| G | 5690 hCoV-19/England/LIVE-A9E5A/2020\|EPI_ISL_517370\|202 | ···················· | ···················· | ···················· | ···················· | ···················· | NNNNNNNNNNNNNNNNNNNN | 1 |
| G | 5691 hCoV-19/England/LIVE-A9E69/2020\|EPI_ISL_517371\|202 | ···················· | ···················· | ···················· | ···················· | ·······NNNNNNNNNNNNN | ···················· | 1 |
| G | 5737 hCoV-19/Australia/VIC5914/2020\|EPI_ISL_518693\|2020 | ···················· | ···················· | ···················· | ···················· | NNNNNNNNNNNNNNN····· | ···················· | 1 |
| G | 5889 hCoV-19/England/LIVE-A840E/2020\|EPI_ISL_517160\|202 | ···················· | ···················· | ···················· | ···················· | ·······NNNNNNNNNNNNN | ···················· | 1 |
| G | 5890 hCoV-19/England/LIVE-A8468/2020\|EPI_ISL_517165\|202 | ···················· | ···················· | ···················· | ···················· | ·······NNNNNNNNNNNNN | ···················· | 1 |
| G | 5891 hCoV-19/England/LIVE-A8477/2020\|EPI_ISL_517166\|202 | ···················· | ···················· | ···················· | ···················· | ·······NNNNNNNNNNNNN | ···················· | 1 |
| G | 5896 hCoV-19/England/LIVE-A850B/2020\|EPI_ISL_517174\|202 | ···················· | ···················· | ···················· | ···················· | ·······NNNNNNNNNNNNN | ···················· | 1 |
| G | 5905 hCoV-19/England/LIVE-A85CF/2020\|EPI_ISL_517186\|202 | ···················· | ···················· | ···················· | ···················· | ·······NNNNNNNNNNNNN | ···················· | 1 |
| G | 6049 hCoV-19/Colombia/I30/2020\|EPI_ISL_526959\|2020-04-1 | ···················· | ···················· | ···················· | ···················· | NNNNNNNNNNNNNNNNNNNN | NNNNNNNNNNNNNNNNNNNN | 1 |
| G | 6054 hCoV-19/Colombia/DC-INS-197/2020\|EPI_ISL_526971\|20 | ···················· | ···················· | ···················· | ···················· | NNNNNNNNNNNNNNNNNNNN | NNNNNNNNNNNNNNNNNNNN | 1 |
| G | 6146 hCoV-19/England/LIVE-A919D/2020\|EPI_ISL_517271\|202 | ···················· | ···················· | ···················· | ···················· | ·······NNNNNNNNNNNNN | ···················· | 1 |
| G | 6170 hCoV-19/Australia/VIC5717/2020\|EPI_ISL_518519\|2020 | ···················· | ···················· | ···················· | ···················· | ···················· | NNNNNNNNNNNNNNNNNNNN | 1 |
| G | 6241 hCoV-19/Australia/VIC3095/2020\|EPI_ISL_520802\|2020 | ···················· | ···················· | ···················· | ···················· | NNNNNNNNNNNNNNNNNNNN | ···················· | 1 |
| G | 6242 hCoV-19/Australia/VIC3096/2020\|EPI_ISL_520803\|2020 | ···················· | ···················· | ···················· | ···················· | NNNNNNNNNNNNNNNNNNNN | ···················· | 1 |
| G | 6249 hCoV-19/Australia/VIC3104/2020\|EPI_ISL_520811\|2020 | ···················· | ···················· | ···················· | ···················· | NNNN················ | ···················· | 1 |
| G | 6269 hCoV-19/Australia/VIC3125/2020\|EPI_ISL_520832\|2020 | ···················· | ···················· | ···················· | ···················· | NNNNNNNNNNNN········ | ···················· | 1 |
| G | 6276 hCoV-19/Australia/VIC3133/2020\|EPI_ISL_520840\|2020 | ···················· | ···················· | ···················· | ···················· | NNNNNNNNNNNNNNNNNNNN | NNNNNNNNNNNNNNNNNNNN | 1 |
| G | 6315 hCoV-19/mink/Netherlands/NB-EMC-25-12/2020\|EPI_ISL | ···················· | ···················· | NNNNNNNNNNNNNNNNNNNN | NNNNNNNNNNNNNNNNNNNN | ···················· | ···················· | 1 |
| G | 6362 hCoV-19/Colombia/AMA-INS-105142/2020\|EPI_ISL_52695 | ···················· | ···················· | ···················· | ···················· | NNNNNNNNNNNNNNNNNNNN | NNNNNNNNNNNNNNNNNNNN | 1 |
| G | 6365 hCoV-19/Colombia/NAR-INS-103313/2020\|EPI_ISL_52695 | ···················· | ···················· | ···················· | ···················· | NNNNNNNNNNNNNNNNNNNN | NNNNNNNNNNNNNNNNNNNN | 1 |
| G | 6401 hCoV-19/England/LIVE-A86CC/2020\|EPI_ISL_517200\|202 | ···················· | ···················· | ···················· | ···················· | ·······NNNNNNNNNNNNN | NNNNNNNNNNNNNNNNNN·· | 1 |
| G | 6402 hCoV-19/England/LIVE-A86EA/2020\|EPI_ISL_517201\|202 | ···················· | ···················· | ···················· | ···················· | ·······NNNNNNNNNNNNN | NNNNNNNNNNNNNNNNNNNN | 1 |
| G | 6403 hCoV-19/England/LIVE-A8714/2020\|EPI_ISL_517203\|202 | ···················· | ···················· | ···················· | ···················· | ·······NNNNNNNNNNNNN | ···················· | 1 |
| G | 6404 hCoV-19/England/LIVE-A8723/2020\|EPI_ISL_517204\|202 | ···················· | ···················· | ···················· | ···················· | ·······NNNNNNNNNNNNN | NNNNNNNNNNNNNNNNNNNN | 1 |
| G | 6406 hCoV-19/England/LIVE-A8750/2020\|EPI_ISL_517206\|202 | ···················· | ···················· | ···················· | ···················· | ·······NNNNNNNNNNNNN | NNNNNNNNNNNNNNNNNNNN | 1 |
| G | 6407 hCoV-19/England/LIVE-A876F/2020\|EPI_ISL_517207\|202 | ···················· | ···················· | ···················· | ···················· | ·······NNNNNNNNNNNN· | ···················· | 1 |
| G | 6408 hCoV-19/England/LIVE-A877E/2020\|EPI_ISL_517208\|202 | ···················· | ···················· | ···················· | ···················· | ·······NNNNNNNNNNNNN | NNNNNNNNNNNNNNNNNNNN | 1 |
| G | 6412 hCoV-19/England/LIVE-A87BA/2020\|EPI_ISL_517212\|202 | ···················· | ···················· | ···················· | ···················· | ·······NNNNNNNNNNNNN | ···················· | 1 |
| G | 6413 hCoV-19/England/LIVE-A87D8/2020\|EPI_ISL_517213\|202 | ···················· | ···················· | ···················· | ···················· | ·······NNNNNNNNNNNNN | ···················· | 1 |
| G | 6417 hCoV-19/England/LIVE-A8811/2020\|EPI_ISL_517217\|202 | ···················· | ···················· | ···················· | ···················· | ·······NNNNNNNNNNNNN | ···················· | 1 |
| G | 6425 hCoV-19/England/LIVE-A8899/2020\|EPI_ISL_517225\|202 | ···················· | ···················· | ···················· | ···················· | ···················· | NNNNNNNNNNNNNNNNNNNN | 1 |
| G | 6428 hCoV-19/England/LIVE-A8EF1/2020\|EPI_ISL_517231\|202 | ···················· | ···················· | ···················· | ···················· | ·······NNNNNNNNNNNN· | ···················· | 1 |
| G | 6430 hCoV-19/England/LIVE-A8F1C/2020\|EPI_ISL_517233\|202 | ···················· | ···················· | ···················· | ···················· | ·······NNNNNNNNNNNNN | ···················· | 1 |
| G | 6432 hCoV-19/England/LIVE-A8F49/2020\|EPI_ISL_517235\|202 | ···················· | ···················· | ···················· | ···················· | ·······NNNNNNNNNNNNN | NNNNNNNNNNNNNNNNNNNN | 1 |
| G | 6440 hCoV-19/England/LIVE-A8FD0/2020\|EPI_ISL_517244\|202 | ···················· | ···················· | ···················· | ···················· | ·······NNNNNNNNNNNNN | ···················· | 1 |
| G | 6442 hCoV-19/England/LIVE-A9009/2020\|EPI_ISL_517247\|202 | ···················· | ···················· | ···················· | ···················· | ·······NNNNNNNNNNNNN | ···················· | 1 |
| G | 6443 hCoV-19/England/LIVE-A9018/2020\|EPI_ISL_517248\|202 | ···················· | ···················· | ···················· | ···················· | ·······NNNNNNNNNNNNN | ···················· | 1 |
| G | 6444 hCoV-19/England/LIVE-A9027/2020\|EPI_ISL_517249\|202 | ···················· | ···················· | ···················· | ···················· | ·······NNNNNNNNNNNNN | ···················· | 1 |
| G | 6455 hCoV-19/England/LIVE-A90DC/2020\|EPI_ISL_517260\|202 | ···················· | ···················· | ···················· | ···················· | ·······NNNNNNNNNNNNN | ···················· | 1 |
| G | 6460 hCoV-19/England/LIVE-A9133/2020\|EPI_ISL_517265\|202 | ···················· | ···················· | ···················· | ···················· | ·······NNNNNNNNNNNNN | ···················· | 1 |
| G | 6569 hCoV-19/Australia/VIC3138/2020\|EPI_ISL_520845\|2020 | ···················· | ···················· | ···················· | ···················· | ·NNNNNNNNNNN········ | ···················· | 1 |
| G | 6576 hCoV-19/Australia/VIC3145/2020\|EPI_ISL_520852\|2020 | ···················· | ···················· | ···················· | ···················· | NNNNNNNNNNNNNNNNNNNN | ···················· | 1 |
| G | 6578 hCoV-19/Australia/VIC3148/2020\|EPI_ISL_520854\|2020 | ···················· | ···················· | ···················· | ···················· | NNNNNNNNNNNNNNNNNNNN | NNNNNNNNNNNNNNNNNNNN | 1 |
| G | 6581 hCoV-19/Australia/VIC3151/2020\|EPI_ISL_520857\|2020 | ···················· | ···················· | ···················· | ···················· | NNNNNNNNNNNNNNNNNNNN | NNNNNNNNNNNNNNNNNNNN | 1 |
| G | 6618 hCoV-19/Australia/VIC3208/2020\|EPI_ISL_520894\|2020 | ···················· | ···················· | ···················· | ···················· | NNNNNNNNNNNNNN······ | ···················· | 1 |
| G | 6657 hCoV-19/England/LIVE-A7593/2020\|EPI_ISL_517040\|202 | ···················· | ···················· | ···················· | ···················· | ·······NNNNNNNNNNNNN | ···················· | 1 |
| G | 6658 hCoV-19/England/LIVE-A75A2/2020\|EPI_ISL_517041\|202 | ···················· | ···················· | ···················· | ···················· | ·······NNNNNNNNNNNNN | NNNNNNNNNNNNNNNNNNNN | 1 |
| G | 6659 hCoV-19/England/LIVE-A75DF/2020\|EPI_ISL_517044\|202 | ···················· | ···················· | ···················· | ···················· | ·······NNNNNNNNNNNNN | ···················· | 1 |
| G | 6660 hCoV-19/England/LIVE-A75EE/2020\|EPI_ISL_517045\|202 | ···················· | ···················· | ···················· | ···················· | ·······NNNNNNNNNNNNN | NNNNNNNNNNNNNNNNNNNN | 1 |
| G | 6663 hCoV-19/England/LIVE-A7627/2020\|EPI_ISL_517049\|202 | ···················· | ···················· | ···················· | ···················· | ·······NNNNNNNNNNNNN | ···················· | 1 |
| G | 6664 hCoV-19/England/LIVE-A7645/2020\|EPI_ISL_517050\|202 | ···················· | ···················· | ···················· | ···················· | ·······NNNNNNNNNNNNN | NNNNNNNNNNNNNNNNNNNN | 1 |
| G | 6665 hCoV-19/England/LIVE-A7663/2020\|EPI_ISL_517051\|202 | ···················· | ···················· | ···················· | ···················· | ·······NNNNNNNNNNNNN | ···················· | 1 |
| G | 6666 hCoV-19/England/LIVE-A7672/2020\|EPI_ISL_517052\|202 | ···················· | ···················· | ···················· | ···················· | ·······NNNNNNNNNNNNN | NNNNNNNNNNNNNNNNNNNN | 1 |
| G | 6671 hCoV-19/England/LIVE-A76FA/2020\|EPI_ISL_517060\|202 | ···················· | ···················· | ···················· | ···················· | ·······NNNNNNNNNNNNN | NNNNNNNNNNNNNNNNNNNN | 1 |
| G | 6675 hCoV-19/England/LIVE-A7760/2020\|EPI_ISL_517066\|202 | ···················· | ···················· | ···················· | ···················· | ·······NNNNNNNNNNNNN | NNNNNNNNNNNNNNNNNNNN | 1 |
| G | 6679 hCoV-19/England/LIVE-A77D9/2020\|EPI_ISL_517072\|202 | ···················· | ···················· | ···················· | ···················· | ·······NNNNNNNNNNNNN | NNNNNNNNNNNNNNNNNNNN | 1 |
| G | 6680 hCoV-19/England/LIVE-A77E8/2020\|EPI_ISL_517073\|202 | ···················· | ···················· | ···················· | ···················· | ·······NNNNNNNNNNNNN | ···················· | 1 |
| G | 6684 hCoV-19/England/LIVE-A7821/2020\|EPI_ISL_517077\|202 | ···················· | ···················· | ···················· | ···················· | ·······NNNNNNNNNNNNN | ···················· | 1 |
| G | 6685 hCoV-19/England/LIVE-A7830/2020\|EPI_ISL_517078\|202 | ···················· | ···················· | ···················· | ···················· | ·······NNNNNNNNNNNNN | ···················· | 1 |
| G | 6691 hCoV-19/England/LIVE-A78A9/2020\|EPI_ISL_517084\|202 | ···················· | ···················· | ···················· | ···················· | ·······NNNNNNNNNNNNN | ···················· | 1 |
| G | 6692 hCoV-19/England/LIVE-A78B8/2020\|EPI_ISL_517085\|202 | ···················· | ···················· | ···················· | ···················· | ·······NNNNNNNNNNNNN | NNNNNNNNNNNNNNNNNNNN | 1 |

G 6694 hCoV-19/England/LIVE-A7F3B/2020|EPI_ISL_517093|202 ···················· ···················· ···················· ···················· ·······NNNNNNNNNNNNN NNNNNNNNNNNNNNNNNNNN 1
G 6695 hCoV-19/England/LIVE-A7F68/2020|EPI_ISL_517095|202 ···················· ···················· ···················· ···················· ·······NNNNNNNNNNNNN NNNNNNNNNNNNNNNNNNNN 1
G 6777 hCoV-19/USA/VA-DCLS-0586/2020|EPI_ISL_526807|2020- ···················· ···················· ···················· ···················· ·······NNNNNNNNNNNNN NNNNNNNNNNNNNNNNNNNN 1
G 6853 hCoV-19/USA/SC-DHEC-0535/2020|EPI_ISL_529083|2020- ···················· ···················· ···················· ···················· ·NNNNNNNN··········· ···················· 1
G 6917 hCoV-19/Australia/VIC5585/2020|EPI_ISL_518402|2020 ···················· ···················· ···················· ···················· NNNNNNNNNNNNNNNNNNNN ···················· 1
G 7167 hCoV-19/Switzerland/BS-UHB-42185807/2020|EPI_ISL_5 ···················· ···················· ···················· ···················· NNNNNNNNNNNNNNNNNNNN NNNNNNNNNNNNNNNNNNNN 1
G 7169 hCoV-19/England/LIVE-A8064/2020|EPI_ISL_517108|202 ···················· ···················· ···················· ···················· ·······NNNNNNNNNNNNN NNNNNNNNNNNNNNNNNNNN 1
G 7170 hCoV-19/England/LIVE-A80FB/2020|EPI_ISL_517116|202 ···················· ···················· ···················· ···················· ·······NNNNNNNNNNNNN NNNNNNNNNNNNNNNNNNNN 1
G 7171 hCoV-19/England/LIVE-A8161/2020|EPI_ISL_517121|202 ···················· ···················· ···················· ···················· ···················· NNNNNNNNNNNNNNNNNNNN 1
G 7172 hCoV-19/England/LIVE-A818F/2020|EPI_ISL_517123|202 ···················· ···················· ···················· ···················· ·······NNNNNNNNNNNNN NNNNNNNNNNNNNNNNNNNN 1
G 7176 hCoV-19/England/LIVE-A8310/2020|EPI_ISL_517146|202 ···················· ···················· ···················· ···················· ·······NNNNNNNNNNNNN ···················· 1
G 7183 hCoV-19/Australia/VIC5609/2020|EPI_ISL_518424|2020 ···················· ···················· ···················· ···················· NNNNNNNNNNNNNNNNNNNN ···················· 1
G 7322 hCoV-19/Australia/VIC3048/2020|EPI_ISL_520761|2020 ···················· ···················· ···················· ···················· NNNNNNNNNNNNNNNNNNNN NNNNNNNNNK·········· 1
G 7326 hCoV-19/Australia/VIC3060/2020|EPI_ISL_520767|2020 ···················· ···················· ···················· ···················· NNNN················ ···················· 1
G 7330 hCoV-19/Australia/VIC3064/2020|EPI_ISL_520771|2020 ···················· ···················· ···················· ···················· NNNNNNNNNNNNNN······ ···················· 1
G 7332 hCoV-19/Australia/VIC3066/2020|EPI_ISL_520773|2020 ···················· ···················· ···················· ···················· NNNNNNNNNNNNNNNNNNNN ···················· 1
G 7335 hCoV-19/Australia/VIC3070/2020|EPI_ISL_520777|2020 ···················· ···················· ···················· ···················· NNNNNNNNNNNNNNNNNNNN ···················· 1
G 7341 hCoV-19/Australia/VIC3076/2020|EPI_ISL_520783|2020 ···················· ···················· ···················· ···················· ··NNNNNNNNNNNNNNNNNN ···················· 1
G 7350 hCoV-19/Australia/VIC3085/2020|EPI_ISL_520792|2020 ···················· ···················· ···················· ···················· NNNN················ ···················· 1
G 7429 hCoV-19/USA/FL-BPHL-0914/2020|EPI_ISL_517804|2020- ···················· ···················· ···················· ···················· NNNNNNNNNNNNNNNNNNNN ···················· 1
G 7431 hCoV-19/USA/FL-BPHL-0916/2020|EPI_ISL_517806|2020- ···················· ···················· ···················· ···················· NNNNNNNNNNNNNNNNNNNN NNNNNNNNNNNNNNN····· 1
G 7435 hCoV-19/USA/FL-BPHL-0920/2020|EPI_ISL_517810|2020- ···················· ···················· ···················· ···················· ···················· NNNNNNNNNNNNNNNNNNNN 1
G 7442 hCoV-19/USA/FL-BPHL-0929/2020|EPI_ISL_517819|2020- ···················· ···················· ···················· ···················· NNNNNNNNNNNNNNNNNNNN ···················· 1
G 7443 hCoV-19/USA/FL-BPHL-0931/2020|EPI_ISL_517821|2020- ···················· ···················· ···················· ···················· NNNNNNNNNNNNNNNNNNNN NNNNNNNNNNNNNNNNNNNN 1
G 7448 hCoV-19/USA/FL-BPHL-0939/2020|EPI_ISL_517829|2020- ···················· ···················· ···················· ···················· NNNNNNNNNNNNNNNNNNNN NNNNNNNNNNNNNNNNNNNN 1
G 7449 hCoV-19/USA/FL-BPHL-0940/2020|EPI_ISL_517830|2020- ···················· ···················· ···················· ···················· NNN················· NNNNNNNNNNNNNNNNNNNN 1
G 7453 hCoV-19/USA/FL-BPHL-0944/2020|EPI_ISL_517834|2020- ···················· ···················· ···················· ···················· NNNNNNNNNNNNNNNNNNNN NNNNNNNNNNNNNNNNNNNN 1
G 7459 hCoV-19/USA/FL-BPHL-1002/2020|EPI_ISL_517892|2020- ···················· ···················· ···················· ···················· NNNNNNNNNNNNNNNNNNNN ···················· 1
G 7460 hCoV-19/USA/FL-BPHL-1003/2020|EPI_ISL_517893|2020- ···················· ···················· ···················· ···················· NNNNNNNNNNNNNNNNNNNN NNNNNNNNNNNNNNNNNNNN 1
G 7461 hCoV-19/USA/FL-BPHL-1004/2020|EPI_ISL_517894|2020- ···················· ···················· ···················· ···················· NNNNNNNNNNNNNNNNNNNN NNNNNNNNNNNNNNNNNNNN 1
G 7495 hCoV-19/Australia/VIC3367/2020|EPI_ISL_521046|2020 ···················· ···················· ···················· ···················· NNNNNNNNNNNNNNNNNNNN ···················· 1
G 7537 hCoV-19/Australia/VIC3415/2020|EPI_ISL_521089|2020 ···················· ···················· ···················· ···················· ········NNNNNNNNNNNN ···················· 1
G 7539 hCoV-19/Australia/VIC3418/2020|EPI_ISL_521092|2020 ···················· ···················· ···················· ···················· ··················NN NNNNNNNNNNNNNNNNNNNN 1
G 7542 hCoV-19/Australia/VIC3424/2020|EPI_ISL_521095|2020 ···················· ···················· ···················· ···················· NNNNNNNNNNNNNNNNNNNN NNNNNNNNNNNNNNNNNNNN 1
G 7544 hCoV-19/Australia/VIC3427/2020|EPI_ISL_521097|2020 ···················· ···················· ···················· ···················· NNNNNNNNNNNNNNNNNNNN NNNNNNNNNNNNNNNNNNNN 1
G 7545 hCoV-19/Australia/VIC3428/2020|EPI_ISL_521098|2020 ···················· ···················· ···················· ···················· NNNN················ ···················· 1
G 7680 hCoV-19/USA/MI-MDHHS-SC21770/2020|EPI_ISL_529888|2 ···················· ·············A······ ···················· ···················· ·NNNNN·NNN····NN···· ···················· 1
G 7684 hCoV-19/USA/FL-BPHL-0952/2020|EPI_ISL_517842|2020- ···················· ···················· ···················· ···················· NNNNNNNNNNNNNNNNNNNN ···················· 1
G 7685 hCoV-19/USA/FL-BPHL-0953/2020|EPI_ISL_517843|2020- ···················· ···················· ···················· ···················· NNNNNNNNNNNNNNNNNNNN NNNNNNNNNNNNNNNNNNNN 1
G 7686 hCoV-19/USA/FL-BPHL-0954/2020|EPI_ISL_517844|2020- ···················· ···················· ···················· ···················· NNNN············NNNN NNN·············NN·· 1
G 7697 hCoV-19/USA/FL-BPHL-0971/2020|EPI_ISL_517861|2020- ···················· ···················· ···················· ···················· NNNNNNNNNNNNNNNNNNNN NNNNNNNNNNNNNNNNNNNN 1
G 7699 hCoV-19/USA/FL-BPHL-0973/2020|EPI_ISL_517863|2020- ···················· ···················· ···················· ···················· NNNNNNNNNNNNNNNNNNNN NNNNNNNNNNNNNN······ 1
G 7702 hCoV-19/USA/FL-BPHL-0976/2020|EPI_ISL_517866|2020- ···················· ···················· ···················· ···················· NNNNNNNNNNNNNNNNNNNN NNNNNNNNNNNNNNNNNNNN 1
G 7706 hCoV-19/USA/FL-BPHL-0980/2020|EPI_ISL_517870|2020- ···················· ···················· ···················· ···················· ·NNNNNNNNNNNNNNNNNNN NNNNNNNNNNNNNNNNNNNN 1
G 7707 hCoV-19/USA/FL-BPHL-0981/2020|EPI_ISL_517871|2020- ···················· ···················· ···················· ···················· NNNNNNNNNNNNNNNNNNNN ···················· 1
G 7710 hCoV-19/USA/FL-BPHL-0987/2020|EPI_ISL_517877|2020- ···················· ···················· ···················· ···················· ·NNNNNNNNNNNNN······ ···················· 1
G 7716 hCoV-19/USA/FL-BPHL-0999/2020|EPI_ISL_517889|2020- ···················· ·············T······ ···················· ···················· ·NN················N NNNNNNNNNNNNNNNNNNNN 1
G 7718 hCoV-19/Australia/VIC2506/2020|EPI_ISL_522201|2020 ···················· ···················· ···················· ···················· NNNN················ ···················· 1
G 7733 hCoV-19/Australia/VIC2522/2020|EPI_ISL_522216|2020 ···················· ···················· ···················· ···················· NNNNNNNNNNNNNNNNNNNN ···················· 1
G 7738 hCoV-19/Australia/VIC2528/2020|EPI_ISL_522221|2020 ···················· ···················· ···················· ···················· NNNNNNNNNNNNNNNNNNNN ···················· 1
G 7749 hCoV-19/Australia/VIC2573/2020|EPI_ISL_522251|2020 ···················· ···················· ···················· ···················· NNNNNNNNNNNNNNNNNNNN NNNNNNNNNNNNNNNNNNNN 1
G 7774 hCoV-19/Mexico/CMX-INMEGEN-22/2020|EPI_ISL_523500| ···················· ···················· ···················· ···················· NNNNNNNNNNNNNNNNNNNN NNNNNNNNNNNNNNNNNNNN 1
G 7933 hCoV-19/USA/MI-MDHHS-SC21699/2020|EPI_ISL_529818|2 ···················· ···················· ···················· ···················· NNNNNNNNNNNNNNNNNNNN NNNNNNNNNNNNNNNNNNNN 1
G 7935 hCoV-19/USA/MI-MDHHS-SC21713/2020|EPI_ISL_529832|2 ···················· ···················· ···················· ···················· NNNNNNNNNNNNNNNNNNNN NNNNNNNNNNNNNNNNNNNN 1
G 7936 hCoV-19/USA/MI-MDHHS-SC21731/2020|EPI_ISL_529850|2 ···················· ···················· ···················· ···················· NNNNNNNNNNNNNNNNNNNN NNNNNNNNNNNNNNNNNNNN 1
G 7945 hCoV-19/USA/FL-BPHL-0907/2020|EPI_ISL_517797|2020- ···················· ···················· ···················· ···················· NNNNNNNNNNNNNNNNNNNN NNNNNNNNNNNNNNNNNNNN 1
G 7946 hCoV-19/USA/FL-BPHL-0908/2020|EPI_ISL_517798|2020- ···················· ···················· ···················· ···················· NNNNNNNNNNNNNNNNNNNN NNNNNNNNNNNNNNNNNNNN 1
G 8090 hCoV-19/Ireland/LK-NVRL-72IRL16602/2020|EPI_ISL_52 ···················· ·A·················· ···················· ···················· NNNNNNNNNNNNNNNNNNNN NNNNNNNNNNNNNNNNNNNN 1
G 8114 hCoV-19/Wales/PHWC-169F07/2020|EPI_ISL_529711|2020 ···················· ······C············· ···················· ···················· NNNNNNNNNNNNNN······ ···················· 1
G 8118 hCoV-19/Wales/PHWC-169F70/2020|EPI_ISL_529715|2020 ···················· ···················· ···················· ···················· NNNNNNNNNNNNNNNNNNNN ···················· 1
G 8223 hCoV-19/Australia/VIC2395/2020|EPI_ISL_522112|2020 ···················· ···················· ···················· ···················· NNNNNNNNNNNNNNNNNNNN ···················· 1
G 8228 hCoV-19/Australia/VIC2401/2020|EPI_ISL_522117|2020 ···················· ···················· ···················· ···················· ·NNNNNNNNNNNNNNNNNNN ···················· 1
G 8294 hCoV-19/Australia/VIC2483/2020|EPI_ISL_522185|2020 ···················· ···················· ···················· ···················· NNNNN··············· ···················· 1
G 8615 hCoV-19/Switzerland/BL-UHB-42202242/2020|EPI_ISL_5 ···················· ···················· ···················· ···················· NNNNNNNNNNNNNNNNNNN· ···················· 1
G 8627 hCoV-19/Switzerland/BL-UHB-42209566/2020|EPI_ISL_5 ···················· ···················· ···················· ···················· NNNNNNNNNNNNNNNNNNNN ···················· 1
G 8702 hCoV-19/Wales/PHWC-169C79/2020|EPI_ISL_529697|2020 ···················· ···················· ···················· ·············A······ NNNNNNNNNNNNNNNNNNNN NNNNNN·············· 1
G 8712 hCoV-19/Australia/VIC2329/2020|EPI_ISL_522050|2020 ···················· ···················· ···················· ···················· NNNNNNNNNNNNNNNNNNNN NNNNNNNNNNNNNNNNNNNN 1
G 8714 hCoV-19/Australia/VIC2331/2020|EPI_ISL_522052|2020 ···················· ···················· ···················· ···················· NNNNNNNNNNNNNNNNNNNN NNNNNNNNNNNNNNNNNNNN 1
G 8914 hCoV-19/Switzerland/BS-UHB-42202622/2020|EPI_ISL_5 ···················· ···················· ···················· ···················· NNNNNNNNNNNNNNNNNNNN NNNNNNNNNNNNNNNNNNNN 1
G 8927 hCoV-19/Switzerland/BS-UHB-42203425/2020|EPI_ISL_5 ···················· ···················· ···················· ···················· ·NNNNNNNNNNNNNNNN··· ···················· 1
G 9029 hCoV-19/Australia/VIC2278/2020|EPI_ISL_522000|2020 ···················· ···················· ···················· ···················· NNNNNNNNNNNNNNNNNNNN NNNNNNNNNNNNNNNNNNNN 1
G 9041 hCoV-19/Australia/VIC2290/2020|EPI_ISL_522012|2020 ···················· ···················· ···················· ···················· ···················· NNNN················ 1
G 9049 hCoV-19/Australia/VIC2299/2020|EPI_ISL_522020|2020 ···················· ···················· ···················· ···················· ··NNNNNNNNNNNNNNNNNN NNNNNNNNNNNNNNNNNNNN 1
G 9051 hCoV-19/Australia/VIC2301/2020|EPI_ISL_522022|2020 ···················· ···················· ···················· ···················· NNNNNNNNNNNNNNNNNNNN NNNNNNNNNNNNNNNNNNNN 1
G 9053 hCoV-19/Australia/VIC2303/2020|EPI_ISL_522024|2020 ···················· ···················· ···················· ···················· NNNNNNNNNNNNNNNNNNNN ···················· 1
G 9057 hCoV-19/Australia/VIC2307/2020|EPI_ISL_522028|2020 ···················· ···················· ···················· ···················· NNNNNNNNNNNNNNNN···· ···················· 1
G 9060 hCoV-19/Australia/VIC2310/2020|EPI_ISL_522031|2020 ···················· ···················· ···················· ···················· NNNNNNNNNNNNNNNNNNN· ··K················· 1
G 9061 hCoV-19/Australia/VIC2311/2020|EPI_ISL_522032|2020 ···················· ···················· ···················· ···················· NNNNNNNNNNNNNNNNNNNN ···················· 1
G 9062 hCoV-19/Australia/VIC2312/2020|EPI_ISL_522033|2020 ···················· ···················· ···················· ···················· ·NNN····NNNNNNNNNNNN ···················· 1
G 9063 hCoV-19/Australia/VIC2313/2020|EPI_ISL_522034|2020 ···················· ···················· ···················· ···················· NNNNNNNNNNNNNNNNNNNN ···················· 1

| | | | | | | | | | |
|---|---|---|---|---|---|---|---|---|---|
| G | 9068 | hCoV-19/Australia/VIC2318/2020\|EPI_ISL_522039\|2020 | ···················· | ···················· | ···················· | ···················· | NNNNNNNNNNNNNNNNNNNN | NNNNNNNNNNNNNNNNNNNN | 1 |
| G | 9069 | hCoV-19/Australia/VIC2319/2020\|EPI_ISL_522040\|2020 | ···················· | ···················· | ···················· | ···················· | ··············NNNNNN | ···················· | 1 |
| G | 9078 | hCoV-19/Australia/VIC2328/2020\|EPI_ISL_522049\|2020 | ···················· | ···················· | ···················· | ···················· | NNNNNNNNNNNNNNNNNNNN | NNNNNNNNNNNNNNNNNNNN | 1 |
| G | 9250 | hCoV-19/Wales/PHWC-169B7C/2020\|EPI_ISL_517597\|2020 | ···················· | ···················· | ···················· | ···················· | NNNNNNNNNNNNNNNNNNNN | ···················· | 1 |
| G | 9295 | hCoV-19/Australia/VIC2354/2020\|EPI_ISL_522074\|2020 | ···················· | ···················· | ···················· | ···················· | ··NNNNNNNNNNNNNNNNNN | NNNNNNN············· | 1 |
| G | 9301 | hCoV-19/Australia/VIC2361/2020\|EPI_ISL_522081\|2020 | ···················· | ···················· | ···················· | ···················· | ··NNNNNNNNNNNNNNNNNN | NNNNNNNNNNNNNNN····N | 1 |
| G | 9306 | hCoV-19/Australia/VIC2366/2020\|EPI_ISL_522086\|2020 | ···················· | ···················· | ···················· | ···················· | NNNNNNNNNNNNNNNNNNNN | NNNNNNNNNNNNNNNNNNNN | 1 |
| G | 9307 | hCoV-19/Australia/VIC2367/2020\|EPI_ISL_522087\|2020 | ···················· | ···················· | ···················· | ···················· | NNNNNNNNNNNNNNNNNNNN | NNNNNNNNNNNNNNNNNNNN | 1 |
| G | 9316 | hCoV-19/Australia/VIC2379/2020\|EPI_ISL_522099\|2020 | ···················· | ···················· | ···················· | ···················· | NNNNNNNNNNNNNNNNNNNN | NNNNNNNNNNNNNNNNNNNN | 1 |
| G | 9323 | hCoV-19/Switzerland/BS-UHB-42197742/2020\|EPI_ISL_5 | ···················· | NNNNNNNNNNNNNNNNNNNN | ···················· | ···················· | ···················· | ···················· | 1 |
| H | 226 | hCoV-19/England/NORW-EE5BE/2020\|EPI_ISL_540691\|202 | ···················· | ···················· | ···················· | ···················· | NNNNNNNNNNNNNNNNNNNN | NNNNNNNNNNNNNNNNNNNN | 1 |
| H | 231 | hCoV-19/England/NORW-EF9FD/2020\|EPI_ISL_540698\|202 | ···················· | ···················· | ···················· | ···················· | NNNNNNNNNNNNNNNNNNNN | NNNNNNNNNNNNNNNNNNNN | 1 |
| H | 232 | hCoV-19/Spain/MD-IBV-99007126/2020\|EPI_ISL_541900\| | ···················· | ···················· | ···················· | ···················· | NNNNNNNNNNNNNNNNNNNN | NNNNNNNNNNNNNNNNNNNN | 1 |
| H | 233 | hCoV-19/Spain/MD-IBV-99007128/2020\|EPI_ISL_541902\| | ···T················ | ···················· | ···················· | ···················· | NNNNNNNNNNNNNNNNNNNN | NNNNNNNNNNNNNNN··NNN | 1 |
| H | 236 | hCoV-19/Spain/MD-IBV-99007132/2020\|EPI_ISL_541905\| | ···················· | ···················· | ···················· | ···················· | NNNNNNNNNNNNNNNNNNNN | NNNNNNNNNNNNNNNNNNNN | 1 |
| H | 237 | hCoV-19/Spain/MD-IBV-99007133/2020\|EPI_ISL_541906\| | ···················· | ···················· | ···················· | ···················· | NNNNNNNNNNNNNNNNNNNN | NNNNNNNNNNNNNNNNNNNN | 1 |
| H | 238 | hCoV-19/Spain/MD-IBV-99007134/2020\|EPI_ISL_541907\| | ···················· | ···················· | ···················· | ···················· | NNNNNNNNNNNNNNNNNNNN | NNNNNNNNNNNNNNNNNNNN | 1 |
| H | 239 | hCoV-19/Spain/MD-IBV-99007135/2020\|EPI_ISL_541908\| | ···················· | ···················· | ···················· | ···················· | NNNNNNNNNNNNNNNNNNNN | NNNNNNNNNNNNNNNNNNNN | 1 |
| H | 240 | hCoV-19/Spain/MD-IBV-99007138/2020\|EPI_ISL_541911\| | ···················· | ···················· | ···················· | ···················· | NNNNNNNNNNNNNNNNNNNN | NNNNNNNNNNNNNNNNNNNN | 1 |
| H | 241 | hCoV-19/Spain/MD-IBV-99007139/2020\|EPI_ISL_541912\| | ···················· | ···················· | ···················· | ···················· | NNNNNNNNNNNNNNNNNNNN | ··············N····N | 1 |
| H | 242 | hCoV-19/Spain/MD-IBV-99007140/2020\|EPI_ISL_541913\| | ···················· | ···················· | ···················· | ···················· | NNNNNNNNNNNNNNNNNNNN | NNNNNNNNNNNNNNNNNNNN | 1 |
| H | 243 | hCoV-19/Spain/MD-IBV-99007142/2020\|EPI_ISL_541915\| | ···················· | ···················· | ···················· | ···················· | NNNNNNNNNNNNNNNNNNNN | NNNNNNNNNNNNNNNNNNNN | 1 |
| H | 245 | hCoV-19/Spain/MD-IBV-99007145/2020\|EPI_ISL_541917\| | ···················· | ···················· | ···················· | ···················· | NNNNNNNNNNNNNNNNNNNN | NNNNNNNNNNNNNNNNNNNN | 1 |
| H | 246 | hCoV-19/Spain/MD-IBV-99007146/2020\|EPI_ISL_541918\| | ···················· | ···················· | ···················· | ···················· | NNNNNNNNNNNNNNNNNNNN | NNNNNNNNNNNNNNNNNNNN | 1 |
| H | 247 | hCoV-19/Spain/MD-IBV-99007147/2020\|EPI_ISL_541919\| | ···················· | ···················· | ···················· | ···················· | NNNNNNNNNNNNNNNNNNNN | NNNNNNNNNNNNNNNNNNNN | 1 |
| H | 248 | hCoV-19/Spain/MD-IBV-99007148/2020\|EPI_ISL_541920\| | ···················· | ···················· | ···················· | ···················· | NNNNNNNNNNNNNNNNNNNN | NNNNNNNNNNNNNNNNNNNN | 1 |
| H | 249 | hCoV-19/Spain/MD-IBV-99007149/2020\|EPI_ISL_541921\| | ···················· | ···················· | ···················· | ···················· | NNNNNNNNNNNNNNNNNNNN | NNNNNNNNNNNNNNNNNNNN | 1 |
| H | 250 | hCoV-19/Spain/MD-IBV-99007150/2020\|EPI_ISL_541922\| | ···················· | ···················· | ···················· | ···················· | ·NNNNNN············· | ···················· | 1 |
| H | 251 | hCoV-19/Spain/MD-IBV-99007152/2020\|EPI_ISL_541924\| | ···················· | ···················· | ···················· | ···················· | NNNNNNNNNNNNNNNNNNNN | NNNNNNNNNNNNNNNNNNNN | 1 |
| H | 252 | hCoV-19/Spain/MD-IBV-99007153/2020\|EPI_ISL_541925\| | ···················· | ···················· | ···················· | ···················· | NNNNNNNNNNNNNNNNNNNN | NNNNNNNNNNNNNNNNNNNN | 1 |
| H | 253 | hCoV-19/Spain/MD-IBV-99007154/2020\|EPI_ISL_541926\| | ···················· | ···················· | ···················· | ···················· | NNNNNNNNNNNNNNNNNNNN | NNNNNNNNNNNNNNNNNNNN | 1 |
| H | 255 | hCoV-19/Spain/MD-IBV-99007158/2020\|EPI_ISL_541928\| | ···················· | ···················· | ···················· | ···················· | NNNNNNNNNNNNNNNNNNNN | ···················· | 1 |
| H | 256 | hCoV-19/Spain/MD-IBV-99007159/2020\|EPI_ISL_541929\| | ···················· | ···················· | ···················· | ···················· | NNNNNNNNNNNNNNNNNNNN | NNNNNNNNNNNNNNNNNNNN | 1 |
| H | 457 | hCoV-19/England/LIVE-AABF8/2020\|EPI_ISL_540604\|202 | ···················· | ···················· | ··········T········· | ···················· | ······NNNNNNNNNNNNNN | ···················· | 1 |
| H | 458 | hCoV-19/England/LIVE-AAC22/2020\|EPI_ISL_540605\|202 | ···················· | ···················· | ···················· | ···················· | NNNNNNNNNNNNNNNNNNNN | NNNNNNNNNNNNNNNNNNNN | 1 |
| H | 460 | hCoV-19/England/LIVE-AACF5/2020\|EPI_ISL_540613\|202 | ···················· | ···················· | ···················· | ···················· | NNNNNNNNNNNNNNNNNNNN | NNNNNNNNNNNNNNNNNNNN | 1 |
| H | 462 | hCoV-19/England/LIVE-AAD4D/2020\|EPI_ISL_540617\|202 | ···················· | ···················· | ···················· | ···················· | NNNNNNNNNNNNNNNNNNNN | NNNNNNNNNNNNNNNNNNNN | 1 |
| H | 465 | hCoV-19/England/LIVE-AAE2C/2020\|EPI_ISL_540620\|202 | ···················· | ···················· | ···················· | ···················· | NNNNNNNNNNNNNNNNNNNN | NNNNNNNNNNNNNNNNNNNN | 1 |
| H | 469 | hCoV-19/England/LIVE-AAFA1/2020\|EPI_ISL_540625\|202 | ···················· | ···················· | ···················· | ···NN··············· | ······NNNNNNNNNNNNNN | ···················· | 1 |
| H | 497 | hCoV-19/Spain/MD-IBV-99007163/2020\|EPI_ISL_541933\| | ···················· | ···················· | ···················· | ···················· | NNNNNNNNNNNNNNNNNNNN | ···················· | 1 |
| H | 499 | hCoV-19/Spain/MD-IBV-99007166/2020\|EPI_ISL_541936\| | ···················· | ···················· | ···················· | ···················· | NNNNNNNNNNNNNNNNNNNN | NNNNNNNNNNNNNNNNNNNN | 1 |
| H | 500 | hCoV-19/Spain/MD-IBV-99007168/2020\|EPI_ISL_541938\| | ···················· | ···················· | ···················· | ···················· | NNNNNNNNNNNNNNNNNNNN | NNNNNNNNNNNNNNNNNNNN | 1 |
| H | 502 | hCoV-19/Spain/MD-IBV-99007175/2020\|EPI_ISL_541941\| | ···················· | ···················· | ···················· | ···················· | NNNNNNNNNNNNNNNNNNNN | ···················· | 1 |
| H | 503 | hCoV-19/Spain/IB-IBV-99007181/2020\|EPI_ISL_541949\| | ···················· | ···················· | ···················· | ···················· | NNNNNNNNNNNNNNNNNNNN | ···················· | 1 |
| H | 504 | hCoV-19/Spain/IB-IBV-99007182/2020\|EPI_ISL_541950\| | ···················· | ···················· | ···················· | ···················· | NNNNNNNNNNNNNNNNNNNN | NNNNNNNNNNNNNNNNN··· | 1 |
| H | 505 | hCoV-19/Spain/IB-IBV-99007192/2020\|EPI_ISL_541951\| | ···················· | ···················· | ···················· | ···················· | NNNNNNNNNNNNNNNNNNNN | ···················· | 1 |
| H | 506 | hCoV-19/Spain/IB-IBV-99007194/2020\|EPI_ISL_541953\| | ···················· | ···················· | ···················· | ···················· | NNNNNNNNNNNNNNNNNNNN | NNNNNNNNNNNNNNNNNNNN | 1 |
| H | 507 | hCoV-19/Spain/IB-IBV-99007209/2020\|EPI_ISL_541956\| | ···················· | ···················· | ···················· | ···················· | NNNNNNNNNNNNNNNNNNNN | ···················· | 1 |
| H | 508 | hCoV-19/Spain/IB-IBV-99007217/2020\|EPI_ISL_541958\| | ···················· | ···················· | ···················· | ···················· | NNNNNNNNNNNNNNNNNNNN | NNNNNNNNNNNNNNNNNNNN | 1 |
| H | 509 | hCoV-19/Spain/IB-IBV-99007241/2020\|EPI_ISL_541963\| | ···················· | ···················· | ···················· | ···················· | NNNNNNNNNNNNNNNNNNNN | NNNNNNNNNNNNNNNNNNNN | 1 |
| H | 510 | hCoV-19/Spain/IB-IBV-99007245/2020\|EPI_ISL_541964\| | ···················· | ···················· | ···················· | ···················· | NNNNNNNNNNNNNNNNNNNN | NNNNNNNNNNNNNNNNNNNN | 1 |
| H | 511 | hCoV-19/Spain/IB-IBV-99007252/2020\|EPI_ISL_541965\| | ···················· | ···················· | ···················· | ···················· | NNNNNNN············· | ···················· | 1 |
| H | 783 | hCoV-19/England/QEUH-960FAE/2020\|EPI_ISL_532020\|20 | ···················· | ···················· | ···················· | NNNNNNNNNNNNNNNNNNNN | ···················· | ···················· | 1 |
| H | 967 | hCoV-19/England/LIVE-AA967/2020\|EPI_ISL_540588\|202 | ···················· | ···················· | ···················· | ···················· | ······NNNNNNNNNNNNNN | NNNNNNNNNNNNNNNNNNNN | 1 |
| H | 969 | hCoV-19/England/LIVE-AA9D0/2020\|EPI_ISL_540591\|202 | ···················· | ···················· | ···················· | ···················· | NNNNNNNNNNNNNNNNNNNN | ···················· | 1 |
| H | 971 | hCoV-19/England/LIVE-AAA37/2020\|EPI_ISL_540593\|202 | ···················· | ···················· | ···················· | ···················· | ······NNNNNNNNNNNNNN | ···················· | 1 |
| H | 1020 | hCoV-19/Spain/MD-IBV-99007121/2020\|EPI_ISL_541895\| | ···················· | ···················· | ···················· | ···················· | NNNNNNNNNNNNNNNNNNNN | NNNNNNNNNNNNNNNNNNNN | 1 |
| H | 1021 | hCoV-19/Spain/MD-IBV-99007122/2020\|EPI_ISL_541896\| | ···················· | ···················· | ···················· | ···················· | NNNNNNNNNNNNNNNNNNNN | NNNNNNNNNNNNNNNNNNNN | 1 |
| H | 1022 | hCoV-19/Spain/MD-IBV-99007123/2020\|EPI_ISL_541897\| | ···················· | ···················· | ···················· | ···················· | NNNNNNNNNNNNNNNNNNNN | NNNNNNNNNNNNNNNNNNNN | 1 |
| H | 1023 | hCoV-19/Spain/MD-IBV-99007124/2020\|EPI_ISL_541898\| | ···················· | ···················· | ···················· | ···················· | NNNNNNNNNNNNNNNNNNNN | NNNNNNNNNNNNNNNNNNNN | 1 |
| H | 1024 | hCoV-19/Spain/MD-IBV-99007125/2020\|EPI_ISL_541899\| | ···················· | ···················· | ···················· | ···················· | NNNNNNNNNNNNNNNNNNNN | NNNNNNNNNNNNNNNNNNNN | 1 |
| H | 1273 | hCoV-19/Spain/MD-IBV-99007088/2020\|EPI_ISL_541878\| | ···················· | ···················· | ···················· | ···················· | NNNNNNNNNNNNNNNNNNNN | NNNNNNNNNNNNNNNNNNNN | 1 |
| H | 1277 | hCoV-19/Spain/MD-IBV-99007115/2020\|EPI_ISL_541889\| | ···················· | ···················· | ···················· | ···················· | NNNNNNN·NNNN·NNNNNNN | NNNNNNNNNNNNNNNNNNNN | 1 |
| H | 1278 | hCoV-19/Spain/MD-IBV-99007116/2020\|EPI_ISL_541890\| | ···················· | ···················· | ···················· | ···················· | NNNNNNNNNNNNNNNNNNNN | NNNNNNNNNNNNNNNNNNNN | 1 |
| H | 1279 | hCoV-19/Spain/MD-IBV-99007118/2020\|EPI_ISL_541892\| | ···················· | ···················· | ···················· | ···················· | NNNNNNNNNNNNNNNNNNNN | NNNNNNNNNNNNNNNNNNNN | 1 |
| H | 1280 | hCoV-19/Spain/MD-IBV-99007120/2020\|EPI_ISL_541894\| | ···················· | ···················· | ···················· | ···················· | ·NNNNNN·NNNN·NNNNNNN | NNNNNNNNNNNNNNN····· | 1 |
| H | 1348 | hCoV-19/Spain/AN-IBV-98001701/2020\|EPI_ISL_538131\| | ···················· | ···················· | ···················· | ···················· | NNNNNNNNNNNNNNNNNNNN | NNNNNNNNNNNNNNNNNNNN | 1 |
| H | 1349 | hCoV-19/Spain/AN-IBV-98001706/2020\|EPI_ISL_538134\| | ···················· | ···················· | ···················· | ···················· | NNNNNNNNNNNNNNNNNNNN | NNNNNNNNNNNNNNNNNNNN | 1 |
| H | 1350 | hCoV-19/Spain/AN-IBV-98001709/2020\|EPI_ISL_538135\| | ···················· | ···················· | ···················· | ···················· | NNNNNNNNNNNNNNNNNNNN | NNNNNNNNNNNNNNNNNNNN | 1 |
| H | 1353 | hCoV-19/Spain/AN-IBV-98001770/2020\|EPI_ISL_538147\| | ···················· | ···················· | ···················· | ···················· | NNNNNNNNNNNNNNNNNNNN | NNNNNNNNNNNNNNNNNNNN | 1 |
| H | 1354 | hCoV-19/Spain/AN-IBV-98001774/2020\|EPI_ISL_538150\| | ···················· | ···················· | ···················· | ···················· | NNNNNNNNNNNNNNNNNNNN | NNNNNNNNNNNNNNNNNNNN | 1 |
| H | 1357 | hCoV-19/Spain/AN-IBV-98001780/2020\|EPI_ISL_538153\| | ···················· | ···················· | ···················· | ···················· | NNNNNNNNNNNNNNNNNNNN | NNNNNNNNNNNNNNNNNNNN | 1 |
| H | 1358 | hCoV-19/Spain/AN-IBV-98001782/2020\|EPI_ISL_538154\| | ···················· | ···················· | ···················· | ···················· | NNNNNNNNNNNNNNNNNNNN | NNNNNNNNNNNNNNNNNNNN | 1 |
| H | 1360 | hCoV-19/Spain/AN-IBV-98001788/2020\|EPI_ISL_538157\| | ···················· | ···················· | ···················· | ···················· | NNNNNNNNNNNNNNNNNNNN | NNNNNNNNNNNNNNNNNNNN | 1 |
| H | 1361 | hCoV-19/Spain/AN-IBV-98001801/2020\|EPI_ISL_538160\| | ···················· | ···················· | ···················· | ···················· | NNNNNNNNNNNNNNNNNNNN | ···················· | 1 |
| H | 1362 | hCoV-19/Spain/AN-IBV-98001804/2020\|EPI_ISL_538161\| | ···················· | ···················· | ···················· | ···················· | NNNNNNN············· | ···················· | 1 |
| H | 1364 | hCoV-19/Spain/AN-IBV-98001807/2020\|EPI_ISL_538163\| | ···················· | ···················· | ···················· | ···················· | NNNNNNNNNNNN········ | ···················· | 1 |
| H | 1365 | hCoV-19/Spain/AN-IBV-98001819/2020\|EPI_ISL_538165\| | ···················· | ···················· | ···················· | ···················· | NNNNNNNNNNNNNNNNNNNN | ···················· | 1 |
| H | 1366 | hCoV-19/Spain/AN-IBV-98001835/2020\|EPI_ISL_538167\| | ···················· | ···················· | ···················· | ···················· | NNNNNNNNNNNNNNNNNNNN | NNNNNNNNNNNNNNNNNNNN | 1 |
| H | 1482 | hCoV-19/India/MH-NIV-157478/2020\|EPI_ISL_541707\|20 | ···················· | ···················· | ···················· | ·········NNNNNN····· | ···················· | ···················· | 1 |

| | | | | | | | | |
|---|---|---|---|---|---|---|---|---|
| H | 1483 hCoV-19/India/MH-NIV-157939/2020\|EPI_ISL_541708\|20 | ···················· | ···················· | ···················· | ·········NNNNNNNNNNN | ···················· | ···················· | 1 |
| H | 1484 hCoV-19/India/MH-NIV-159610/2020\|EPI_ISL_541710\|20 | ···················· | ···················· | ···················· | NNNNNNNNNNNNNNN····· | ···················· | ···················· | 1 |
| H | 1496 hCoV-19/India/MH-NIV-192922/2020\|EPI_ISL_541724\|20 | ···················· | ···················· | ···················· | ·····NNNNNNNNNN····· | ···················· | ···················· | 1 |
| H | 1498 hCoV-19/India/MH-NIV-57622/2020\|EPI_ISL_541726\|202 | ···················· | ···················· | ···················· | ·········NNNNNN····· | ···················· | ···················· | 1 |
| H | 1499 hCoV-19/India/MH-NIV-57774/2020\|EPI_ISL_541727\|202 | ···················· | ···················· | ···················· | ·········NNNNNNN···· | ···················· | ···················· | 1 |
| H | 1502 hCoV-19/India/MH-NIV-59448/2020\|EPI_ISL_541730\|202 | ···················· | ···················· | ···················· | ······G·AA·GNNNNNNNN | ···················· | ···················· | 1 |
| H | 1510 hCoV-19/India/MH-NIV-74837/2020\|EPI_ISL_541739\|202 | ···················· | ···················· | ···················· | ·········NNNNNN····· | ···················· | ···················· | 1 |
| H | 1513 hCoV-19/India/MH-NIV-80231/2020\|EPI_ISL_541745\|202 | ···················· | ···················· | ···················· | NNNNNNNNNNNNNNNN··C· | ···················· | ···················· | 1 |
| H | 1516 hCoV-19/India/MH-NIV-96428/2020\|EPI_ISL_541748\|202 | ···················· | ···················· | ···················· | ·········NNNNNN····· | ···················· | ···················· | 1 |
| H | 1710 hCoV-19/Spain/AN-IBV-98001631/2020\|EPI_ISL_538119\| | ···················· | ···················· | ···················· | ···················· | NNNNNNNNNNNNNNNNNNNN | ···················· | 1 |
| H | 1711 hCoV-19/Spain/AN-IBV-98001638/2020\|EPI_ISL_538120\| | ···················· | ···················· | ···················· | ···················· | NNNNNNNNNNNNNNNNNNNN | NNNNNNNNNNNNNNNNNNNN | 1 |
| H | 1712 hCoV-19/Spain/AN-IBV-98001643/2020\|EPI_ISL_538121\| | ···················· | ···················· | ···················· | ···················· | NNNNNNNNNNNNNNNNNNNN | NNNNNNNNNNNNNNNNNNNN | 1 |
| H | 1713 hCoV-19/Spain/AN-IBV-98001646/2020\|EPI_ISL_538123\| | ···················· | ···················· | ···················· | ···················· | NNNNNNNNNNNNNNNNNNNN | NNNNNNNNNNNNNNNNNNNN | 1 |
| H | 1714 hCoV-19/Spain/AN-IBV-98001664/2020\|EPI_ISL_538124\| | ···················· | ···················· | ···················· | ···················· | NNNNNNNNNNNNNNNNNNNN | NNNNNNNNNNNNNNNNNNNN | 1 |
| H | 1715 hCoV-19/Spain/AN-IBV-98001680/2020\|EPI_ISL_538125\| | ···················· | ···················· | ···················· | ···················· | NNNNNNNNNNNNNNNNNNNN | ···················· | 1 |
| H | 1898 hCoV-19/Spain/VC-IBV-98003075/2020\|EPI_ISL_538013\| | ···················· | ···················· | ···················· | ···················· | NNNNNNNNNNNNNNNNNNNN | NNNNNNNNNNNNNNNNNNNN | 1 |
| H | 1899 hCoV-19/Spain/VC-IBV-98003076/2020\|EPI_ISL_538014\| | ···················· | ···················· | ···················· | ···················· | NNNNNNNNNNNNNNNNNNNN | NNNNNNNNNNNNNNNNNNNN | 1 |
| H | 1900 hCoV-19/Spain/VC-IBV-98003078/2020\|EPI_ISL_538015\| | ···················· | ···················· | ···················· | ···················· | NNNNNNN············· | ···················· | 1 |
| H | 1901 hCoV-19/Spain/VC-IBV-98003079/2020\|EPI_ISL_538016\| | ···················· | ···················· | ···················· | ···················· | NNNNNNNNNNNNNNNNNNNN | ···················· | 1 |
| H | 1903 hCoV-19/Spain/VC-IBV-98003082/2020\|EPI_ISL_538018\| | ···················· | ···················· | ···················· | ···················· | NNNNNNN·NNNNNNNNNNNN | ···················· | 1 |
| H | 1905 hCoV-19/Spain/VC-IBV-98003084/2020\|EPI_ISL_538020\| | ···················· | ···················· | ···················· | ···················· | NNNNNNNNNNNNNNNNNNNN | NNNNNNNNNNNNNNNNNNNN | 1 |
| H | 1906 hCoV-19/Spain/VC-IBV-98003085/2020\|EPI_ISL_538021\| | ···················· | ···················· | ···················· | ···················· | NNNNNNNNNNNNNNNNNNNN | NNNNNNNNNNNNNNNNNNNN | 1 |
| H | 1907 hCoV-19/Spain/VC-IBV-98003086/2020\|EPI_ISL_538022\| | ···················· | ···················· | ···················· | ···················· | NNNNNNNNNNNNNNNNNNNN | ···················· | 1 |
| H | 1911 hCoV-19/Spain/VC-IBV-98003090/2020\|EPI_ISL_538026\| | ···················· | ···················· | ···················· | ···················· | NNNNNNNNNNNNNNNNNNNN | ···················· | 1 |
| H | 1912 hCoV-19/Spain/VC-IBV-98003091/2020\|EPI_ISL_538027\| | ···················· | ···················· | ···················· | ···················· | NNNNNNNNNNNNNNNNNNNN | ···········NNNN····· | 1 |
| H | 1913 hCoV-19/Spain/VC-IBV-98003092/2020\|EPI_ISL_538028\| | ···················· | ···················· | ···················· | ···················· | NNNNNNN············· | ···················· | 1 |
| H | 1914 hCoV-19/Spain/VC-IBV-98003094/2020\|EPI_ISL_538030\| | ···················· | ···················· | ···················· | ···················· | ·NNNNNN············· | ···················· | 1 |
| H | 1915 hCoV-19/Spain/VC-IBV-98003097/2020\|EPI_ISL_538033\| | ···················· | ···················· | ···················· | ···················· | NNNNNNNNNNNNNNNNNNNN | NNNNNNNNNNNNNNNNNNNN | 1 |
| H | 1916 hCoV-19/Spain/VC-IBV-98003098/2020\|EPI_ISL_538034\| | ···················· | ···················· | ···················· | ···················· | NNNNNNNNNNNNNNNNNNNN | NNNNNNNNNNNNNNNNNNNN | 1 |
| H | 1918 hCoV-19/Spain/VC-IBV-98003100/2020\|EPI_ISL_538036\| | ···················· | ···················· | ···················· | ···················· | ·NNNNNN············· | ···················· | 1 |
| H | 1919 hCoV-19/Spain/VC-IBV-98003101/2020\|EPI_ISL_538037\| | ···················· | ···················· | ···················· | ···················· | NNNNNNNNNNNNN······· | ·········NNNNNN····· | 1 |
| H | 1920 hCoV-19/Spain/VC-IBV-98003102/2020\|EPI_ISL_538038\| | ···················· | ···················· | ···················· | ···················· | NNNNNNN············· | ···················· | 1 |
| H | 1922 hCoV-19/Spain/VC-IBV-98003105/2020\|EPI_ISL_538041\| | ···················· | ···················· | ···················· | ···················· | ·NNNNNN············· | ···················· | 1 |
| H | 1923 hCoV-19/Spain/VC-IBV-98003106/2020\|EPI_ISL_538042\| | ···················· | ···················· | ···················· | ···················· | NNNNNNNNNNNNNNNNNNNN | NNNNNNNNNNNNNNNNNNNN | 1 |
| H | 1927 hCoV-19/Spain/VC-IBV-98003110/2020\|EPI_ISL_538046\| | ···················· | ···················· | ···················· | ···················· | NNNNNNNNNNNNNNNNNNNN | ·················NNN | 1 |
| H | 1928 hCoV-19/Spain/VC-IBV-98003111/2020\|EPI_ISL_538047\| | ···················· | ···················· | ···················· | ···················· | NNNNNNNNNNNNNNNNNNNN | NNNNNNNNNNNNNNNNNNNN | 1 |
| H | 1929 hCoV-19/Spain/VC-IBV-98003112/2020\|EPI_ISL_538048\| | ···················· | ···················· | ···················· | ···················· | NNNNNNNNNNNNNNNNNNNN | ···················· | 1 |
| H | 1930 hCoV-19/Spain/VC-IBV-98003113/2020\|EPI_ISL_538049\| | ···················· | ···················· | ···················· | ···················· | NNNNNNNNNNNNNNNNNNNN | NNNNNNNNNNNNNNNNNNNN | 1 |
| H | 1931 hCoV-19/Spain/VC-IBV-98003114/2020\|EPI_ISL_538050\| | ···················· | ···················· | ···················· | ···················· | NNNNNNN············· | ···················· | 1 |
| H | 2030 hCoV-19/India/MH-NIV-101459/2020\|EPI_ISL_541681\|20 | ···················· | ···················· | ···················· | NNNNNNNNNNNNNNN····· | ···················· | ···················· | 1 |
| H | 2031 hCoV-19/India/MH-NIV-10418/2020\|EPI_ISL_541682\|202 | ···················· | ···················· | ···················· | ·········C·GTAANNNN· | ···················· | ···················· | 1 |
| H | 2032 hCoV-19/India/MH-NIV-10435/2020\|EPI_ISL_541683\|202 | ···················· | ···················· | ···················· | ·········NNNNNN····· | ···················· | ···················· | 1 |
| H | 2034 hCoV-19/India/MH-NIV-113454/2020\|EPI_ISL_541685\|20 | ···················· | ···················· | ···················· | ·········NNNNNN····· | ···················· | ···················· | 1 |
| H | 2037 hCoV-19/India/MH-NIV-121164/2020\|EPI_ISL_541689\|20 | ···················· | ···················· | ···················· | ·········TTNNNNNNNNN | ···················· | ···················· | 1 |
| H | 2039 hCoV-19/India/MH-NIV-123415/2020\|EPI_ISL_541691\|20 | ···················· | ···················· | ···················· | ·········NNNNNN····· | ···················· | ···················· | 1 |
| H | 2128 hCoV-19/Spain/RI-IBV-99005861/2020\|EPI_ISL_537404\| | ···················· | ···················· | ···················· | ···················· | NNNNNNNNNNNNNNNNNNNN | ···················· | 1 |
| H | 2131 hCoV-19/Spain/RI-IBV-99005868/2020\|EPI_ISL_537411\| | ···················· | ···················· | ···················· | ···················· | NNNNNNN············· | ················NNNN | 1 |
| H | 2213 hCoV-19/USA/WI-UW-1474/2020\|EPI_ISL_541606\|2020-09 | ···················· | ···················· | ···················· | ···················· | NNNNNNNNNNNNNNNNNNNN | NNNNNNNNNNNNNNNNNNNN | 1 |
| H | 2214 hCoV-19/USA/WI-UW-1475/2020\|EPI_ISL_541607\|2020-09 | ···················· | ···················· | ···················· | ···················· | NNNNNNNNNNNNNNNNNNNN | NNNNNNNNNNNNNNNNNNNN | 1 |
| H | 2245 hCoV-19/USA/WI-UW-1514/2020\|EPI_ISL_541640\|2020-09 | ···················· | ···················· | ···················· | ···················· | NNNNNNNNNNNNNNNNNNNN | NNNNNNNNNNNNNNNNNNNN | 1 |
| H | 2247 hCoV-19/USA/WI-UW-1516/2020\|EPI_ISL_541642\|2020-09 | ···················· | ···················· | ···················· | ···················· | NNNNNNNNNNNNNNNNNNNN | NNNNNNNNNNNNNNNNNNNN | 1 |
| H | 2250 hCoV-19/USA/WI-UW-1520/2020\|EPI_ISL_541645\|2020-09 | ···················· | ···················· | ···················· | ···················· | NNNNNNNNNNNNNNNNNNNN | NNNNNNNNNNNNNNNNNNNN | 1 |
| H | 2259 hCoV-19/India/MH-NIV-13804/2020\|EPI_ISL_541698\|202 | ···················· | ···················· | ···················· | ·······NNNNNNNN····· | ···················· | ···················· | 1 |
| H | 2296 hCoV-19/USA/TX-HMH-3999/2020\|EPI_ISL_543286\|2020-0 | ···················· | ···················· | ···················· | ···················· | NNNNNNNNNNNNNNNNNNNN | NNNNNNNNNNNNNNNNNNNN | 1 |
| H | 2297 hCoV-19/USA/TX-HMH-4001/2020\|EPI_ISL_543288\|2020-0 | ···················· | ·············A······ | ···················· | ···················· | NNNNNNNNNNNNNNNNNNNN | NNNNNNNNNNNNNNNNNNNN | 1 |
| H | 2298 hCoV-19/USA/TX-HMH-4004/2020\|EPI_ISL_543290\|2020-0 | ···················· | ···················· | ···················· | ···················· | NNNNNNNNNNNNNNNNNNNN | NNNNNNNNNNNNNNNNNNNN | 1 |
| H | 2299 hCoV-19/USA/TX-HMH-4007/2020\|EPI_ISL_543291\|2020-0 | ···················· | ···················· | ···················· | ···················· | NNNNNNNNNNNNNNNNNNNN | NNNNNNNNNNNNNNNNNNNN | 1 |
| H | 2304 hCoV-19/USA/TX-HMH-4017/2020\|EPI_ISL_543299\|2020-0 | ···················· | ···················· | ···················· | ···················· | NNNNNNNNNNNNNNNNNNNN | NNNNNNNNNNNNNNNNNNNN | 1 |
| H | 2464 hCoV-19/Spain/RI-IBV-99005880/2020\|EPI_ISL_537422\| | ···················· | ···················· | ···················· | ···················· | NNNNNNNNNNNNNNNNNNNN | NNNNNNNNNNNNNNN····· | 1 |
| H | 2465 hCoV-19/Spain/RI-IBV-99005882/2020\|EPI_ISL_537424\| | ···················· | ···················· | ···················· | ···················· | NNNNNNNNNNNNNNNNNNNN | NNNNNNNNNNNNNNN····· | 1 |
| H | 2467 hCoV-19/Spain/RI-IBV-99005885/2020\|EPI_ISL_537427\| | ···················· | ···················· | ···················· | ···················· | NNNNNNNNNNNNNNNNNNNN | ···················· | 1 |
| H | 2468 hCoV-19/Spain/RI-IBV-99005886/2020\|EPI_ISL_537428\| | ···················· | ···················· | ···················· | ···················· | NNNNNNNNNNNNNNNNNNNN | NNNNNNNNNNNNNNNNNNNN | 1 |
| H | 2469 hCoV-19/Spain/RI-IBV-99005887/2020\|EPI_ISL_537429\| | ···················· | ···················· | ···················· | ···················· | NNNNNNNNNNNNNNNNNNNN | NNNNNNNNNNNNNNNNNNNN | 1 |
| H | 2471 hCoV-19/Spain/RI-IBV-99005890/2020\|EPI_ISL_537432\| | ···················· | ···················· | ···················· | ···················· | NNNNNNNNNNNNNNNNNNNN | NNNNNNNNNNNNNNNNNNNN | 1 |
| H | 2475 hCoV-19/Spain/RI-IBV-99005899/2020\|EPI_ISL_537438\| | ···················· | ···················· | ···················· | ···················· | NNNNNNNNNNNNNNNNNNNN | NNNNNNNNNNNNNNNNNNNN | 1 |
| H | 2496 hCoV-19/USA/TX-HMH-3797/2020\|EPI_ISL_543201\|2020-0 | ···················· | ···················· | ···················· | ···················· | NNNNNNNNNNNNNNNNNNNN | NNNNNNNNNNNNNNNNNNNN | 1 |
| H | 2497 hCoV-19/USA/TX-HMH-3798/2020\|EPI_ISL_543202\|2020-0 | ···················· | ···················· | ···················· | ···················· | NNNNNNNNNNNNNNNNNNNN | NNNNNNNNNNNNNNNNNNNN | 1 |
| H | 2498 hCoV-19/USA/TX-HMH-3799/2020\|EPI_ISL_543203\|2020-0 | ···················· | ···················· | ···················· | ···················· | NNNNNNNNNNNNNNNNNNNN | NNNNNNNNNNNNNNNNNNNN | 1 |
| H | 2499 hCoV-19/USA/TX-HMH-3800/2020\|EPI_ISL_543204\|2020-0 | ···················· | ···················· | ···················· | ···················· | NNNNNNNNNNNNNNNNNNNN | NNNNNNNNNNNNNNNNNNNN | 1 |
| H | 2500 hCoV-19/USA/TX-HMH-3801/2020\|EPI_ISL_543205\|2020-0 | ···················· | ···················· | ···················· | ···················· | NNNNNNNNNNNNNNNNNNNN | NNNNNNNNNNNNNNNNNNNN | 1 |
| H | 2503 hCoV-19/USA/TX-HMH-3805/2020\|EPI_ISL_543209\|2020-0 | ···················· | ···················· | ···················· | ···················· | NNNNNNNNNNNNNNNNNNNN | NNNNNNNNNNNNNNNNNNNN | 1 |
| H | 2504 hCoV-19/USA/TX-HMH-3806/2020\|EPI_ISL_543210\|2020-0 | ···················· | ···················· | ···················· | ···················· | NNNNNNNNNNNNNNNNNNNN | NNNNNNNNNNNNNNNNNNNN | 1 |
| H | 2505 hCoV-19/USA/TX-HMH-3807/2020\|EPI_ISL_543211\|2020-0 | ···················· | ···················· | ···················· | ···················· | NNNNNNNNNNNNNNNNNNNN | NNNNNNNNNNNNNNNNNNNN | 1 |
| H | 2506 hCoV-19/USA/TX-HMH-3808/2020\|EPI_ISL_543212\|2020-0 | ···················· | ···················· | ···················· | ···················· | NNNNNNNNNNNNNNNNNNNN | NNNNNNNNNNNNNNNNNNNN | 1 |
| H | 2507 hCoV-19/USA/TX-HMH-3809/2020\|EPI_ISL_543213\|2020-0 | ···················· | ···················· | ···················· | ···················· | NNNNNNNNNNNNNNNNNNNN | NNNNNNNNNNNNNNNNNNNN | 1 |
| H | 2509 hCoV-19/USA/TX-HMH-3811/2020\|EPI_ISL_543215\|2020-0 | ···················· | ···················· | ···················· | ···················· | NNNNNNNNNNNNNNNNNNNN | NNNNNNNNNNNNNNNNNNNN | 1 |
| H | 2510 hCoV-19/USA/TX-HMH-3812/2020\|EPI_ISL_543216\|2020-0 | ···················· | ···················· | ···················· | ···················· | NNNNNNNNNNNNNNNNNNNN | NNNNNNNNNNNNNNNNNNNN | 1 |
| H | 2512 hCoV-19/USA/TX-HMH-3814/2020\|EPI_ISL_543218\|2020-0 | ···················· | ···················· | ···················· | ···················· | NNNNNNNNNNNNNNNNNNNN | NNNNNNNNNNNNNNNNNNNN | 1 |
| H | 2514 hCoV-19/USA/TX-HMH-3816/2020\|EPI_ISL_543220\|2020-0 | ···················· | ···················· | ···················· | ···················· | NNNNNNNNNNNNNNNNNNNN | NNNNNNNNNNNNNNNNNNNN | 1 |
| H | 2516 hCoV-19/USA/TX-HMH-3818/2020\|EPI_ISL_543222\|2020-0 | ···················· | ···················· | ···················· | ···················· | NNNNNNNNNNNNNNNNNNNN | NNNNNNNNNNNNNNNNNNNN | 1 |

```
H	2518 hCoV-19/USA/TX-HMH-3820/2020|EPI_ISL_543224|2020-0	····················	············A·······	····················	····················	NNNNNNNNNNNNNNNNNNNN	NNNNNNNNNNNNNNNNNNNN	1
H	2519 hCoV-19/USA/TX-HMH-3821/2020|EPI_ISL_543225|2020-0	····················	····················	····················	····················	NNNNNNNNNNNNNNNNNNNN	NNNNNNNNNNNNNNNNNNNN	1
H	2520 hCoV-19/USA/TX-HMH-3822/2020|EPI_ISL_543226|2020-0	····················	·····A··············	····················	····················	NNNNNNNNNNNNNNNNNNNN	NNNNNNNNNNNNNNNNNNNN	1
H	2522 hCoV-19/USA/TX-HMH-3825/2020|EPI_ISL_543229|2020-0	····················	····················	····················	····················	·············N······	NN·····NN···········	1
H	2524 hCoV-19/USA/TX-HMH-3829/2020|EPI_ISL_543232|2020-0	····················	····················	····················	····················	NNNNNNNNNNNNNNNNNNNN	NNNNNNNNNNNNNNNNNNNN	1
H	2526 hCoV-19/USA/TX-HMH-3831/2020|EPI_ISL_543234|2020-0	····················	····················	····················	····················	NNNNNNNNNNNNNNNNNNNN	NNNNNNNNNNNNNNNNNNNN	1
H	2530 hCoV-19/USA/TX-HMH-3837/2020|EPI_ISL_543240|2020-0	····················	····················	····················	····················	NNNNNNNNNNNNNNNNNNNN	NNNNNNNNNNNNNNNNNNNN	1
H	2532 hCoV-19/USA/TX-HMH-3839/2020|EPI_ISL_543242|2020-0	····················	····················	····················	····················	NNNNNNNNNNNNNNNNNNNN	NNNNNNNNNNNNNNNNNNNN	1
H	2533 hCoV-19/USA/TX-HMH-3840/2020|EPI_ISL_543243|2020-0	····················	····················	····················	····················	NNNNNNNNNNNNNNNNNNNN	NNNNNNNNNNNNNNNNNNNN	1
H	2534 hCoV-19/USA/TX-HMH-3841/2020|EPI_ISL_543244|2020-0	····················	····················	····················	····················	NNNNNNNNNNNNNNNNNNNN	NNNNNNNNNNNNNNNNNNNN	1
H	2535 hCoV-19/USA/TX-HMH-3842/2020|EPI_ISL_543245|2020-0	····················	····················	····················	····················	NNNNNNNNNNNNNNNNNNNN	NNNNNNNNNNNNNNNNNNNN	1
H	2537 hCoV-19/USA/TX-HMH-3844/2020|EPI_ISL_543247|2020-0	····················	············A·······	····················	····················	·NN··········N··N···	NN·N··NNN·NN····N···	1
H	2538 hCoV-19/USA/TX-HMH-3848/2020|EPI_ISL_543250|2020-0	····················	····················	····················	····················	NNNNNNNNNNNNNNNNNNNN	NNNNNNNNNNNNNNNNNNNN	1
H	2539 hCoV-19/USA/TX-HMH-3849/2020|EPI_ISL_543251|2020-0	····················	····················	····················	····················	NNNNNNNNNNNNNNNNNNNN	NNNNNNNNNNNNNNNNNNNN	1
H	2542 hCoV-19/USA/TX-HMH-3853/2020|EPI_ISL_543254|2020-0	····················	····················	····················	····················	NNNNNNNNNNNNNNNNNNNN	NNNNNNNNNNNNNNNNNNNN	1
H	2543 hCoV-19/USA/TX-HMH-3854/2020|EPI_ISL_543255|2020-0	····················	····················	····················	····················	NNNNNNNNNNNNNNNNNNNN	NNNNNNNNNNNNNNNNNNNN	1
H	2556 hCoV-19/USA/TX-HMH-3904/2020|EPI_ISL_543273|2020-0	····················	····················	····················	····················	NNNNNNNNNNNNNNNNNNNN	NNNNNNNNNNNNNNNNNNNN	1
H	2685 hCoV-19/Spain/RI-IBV-99005907/2020|EPI_ISL_537445|	····················	····················	····················	····················	NNNNNNNNNNNNNNNNNNNN	NNNNNNNNNNNNNNNNNNNN	1
H	2686 hCoV-19/Spain/RI-IBV-99005908/2020|EPI_ISL_537446|	····················	····················	····················	····················	·NNNNNN·············	····················	1
H	2688 hCoV-19/Spain/RI-IBV-99005911/2020|EPI_ISL_537449|	····················	····················	····················	····················	NNNNNNNNNNNNNNNNNNNN	····················	1
H	2689 hCoV-19/Spain/RI-IBV-99005913/2020|EPI_ISL_537451|	····················	····················	····················	····················	NNNNNNNNNNNNNNNNNNNN	NNNNNNNNNNNNNNNNNNNN	1
H	2691 hCoV-19/Spain/RI-IBV-99005923/2020|EPI_ISL_537460|	····················	····················	····················	····················	NNNNNNNNNNNNNNNNNNNN	NNNNNNNNNNNNNNNNNNNN	1
H	2692 hCoV-19/Spain/RI-IBV-99005930/2020|EPI_ISL_537465|	····················	·R··················	····················	····················	NNNNNNNNNNNNNNNNNNNN	NNNNNNNNNNNNNNNNNNNN	1
H	2693 hCoV-19/Spain/RI-IBV-99005931/2020|EPI_ISL_537466|	····················	····················	····················	····················	NNNNNNNNNNNNNNNNNNNN	NNNNNNNN············	1
H	2774 hCoV-19/Wales/PHWC-16A522/2020|EPI_ISL_540901|2020	····················	····················	····················	····················	NNNNNNNNNNNN········	····················	1
H	2775 hCoV-19/Wales/PHWC-16A531/2020|EPI_ISL_540902|2020	····················	····················	····················	····················	NNNNNNNNNNNNNN······	····················	1
H	2789 hCoV-19/USA/TX-HMH-3759/2020|EPI_ISL_543170|2020-0	····················	····················	····················	····················	NNNNNNNNNNNNNNNNNNNN	NNNNNNNNNNNNNNNNNNNN	1
H	2790 hCoV-19/USA/TX-HMH-3760/2020|EPI_ISL_543171|2020-0	····················	····················	····················	····················	NNNNNNNNNNNNNNNNNNNN	NNNNNNNNNNNNNNNNNNNN	1
H	2791 hCoV-19/USA/TX-HMH-3762/2020|EPI_ISL_543173|2020-0	····················	····················	····················	····················	NNNNNNNNNNNNNNNNNNNN	NNNNNNNNNNNNNNNNNNNN	1
H	2792 hCoV-19/USA/TX-HMH-3765/2020|EPI_ISL_543176|2020-0	····················	····················	····················	····················	NNNNNNNNNNNNNNNNNNNN	NNNNNNNNNNNNNNNNNNNN	1
H	2793 hCoV-19/USA/TX-HMH-3768/2020|EPI_ISL_543177|2020-0	····················	····················	····················	····················	NNNNNNNNNNNNNNNNNNNN	NNNNNNNNNNNNNNNNNNNN	1
H	2794 hCoV-19/USA/TX-HMH-3769/2020|EPI_ISL_543178|2020-0	····················	····················	····················	····················	NNNNNNNNNNNNNNNNNNNN	NNNNNNNNNNNNNNNNNNNN	1
H	2795 hCoV-19/USA/TX-HMH-3770/2020|EPI_ISL_543179|2020-0	····················	····················	····················	····················	NNNNNNNNNNNNNNNNNNNN	NNNNNNNNNNNNNNNNNNNN	1
H	2796 hCoV-19/USA/TX-HMH-3773/2020|EPI_ISL_543181|2020-0	····················	····················	····················	····················	NNNNNNNNNNNNNNNNNNNN	NNNNNNNNNNNNNNNNNNNN	1
H	2797 hCoV-19/USA/TX-HMH-3774/2020|EPI_ISL_543182|2020-0	····················	····················	····················	····················	NNNNNNNNNNNNNNNNNNNN	NNNNNNNNNNNNNNNNNNNN	1
H	2798 hCoV-19/USA/TX-HMH-3775/2020|EPI_ISL_543183|2020-0	····················	····················	····················	····················	NNNNNNNNNNNNNNNNNNNN	NNNNNNNNNNNNNNNNNNNN	1
H	2799 hCoV-19/USA/TX-HMH-3778/2020|EPI_ISL_543184|2020-0	····················	····················	····················	····················	NNNNNNNNNNNNNNNNNNNN	NNNNNNNNNNNNNNNNNNNN	1
H	2800 hCoV-19/USA/TX-HMH-3780/2020|EPI_ISL_543185|2020-0	····················	····················	····················	····················	NNNNNNNNNNNNNNNNNNNN	NNNNNNNNNNNNNNNNNNNN	1
H	2801 hCoV-19/USA/TX-HMH-3781/2020|EPI_ISL_543186|2020-0	····················	····················	····················	····················	NNNNNNNNNNNNNNNNNNNN	NNNNNNNNNNNNNNNNNNNN	1
H	2802 hCoV-19/USA/TX-HMH-3782/2020|EPI_ISL_543187|2020-0	····················	····················	····················	····················	NNNNNNNNNNNNNNNNNNNN	NNNNNNNNNNNNNNNNNNNN	1
H	2803 hCoV-19/USA/TX-HMH-3783/2020|EPI_ISL_543188|2020-0	····················	····················	····················	····················	NNNNNNNNNNNNNNNNNNNN	NNNNNNNNNNNNNNNNNNNN	1
H	2805 hCoV-19/USA/TX-HMH-4018/2020|EPI_ISL_543300|2020-0	····················	············A·······	····················	····················	NNNNNNNNNNNNNNNNNNNN	NNNNNNNNNNNNNNNNNNNN	1
H	2812 hCoV-19/USA/TX-HMH-4030/2020|EPI_ISL_543310|2020-0	····················	····················	····················	····················	NNNNNNNNNNNNNNNNNNNN	NNNNNNNNNNNNNNNNNNNN	1
H	2815 hCoV-19/USA/TX-HMH-4033/2020|EPI_ISL_543313|2020-0	····················	NNNNNNNNNNNNNNNNNNNN	····················	····················	····················	····················	1
H	2917 hCoV-19/Spain/CL-IBV-97006153/2020|EPI_ISL_537300|	····················	····················	····················	····················	NNNNNNNNNNNNNNNNNNNN	NNNNNNNNNNNNNNNNNNNN	1
H	2918 hCoV-19/Spain/CL-IBV-97006154/2020|EPI_ISL_537301|	····················	····················	····················	····················	·NNNNNN·············	····················	1
H	2919 hCoV-19/Spain/CL-IBV-97006155/2020|EPI_ISL_537302|	····················	····················	····················	····················	NNNNNNNNNNNNNNNNNNNN	NNNNNNNNNNNNNNNNNNNN	1
H	2920 hCoV-19/Spain/CL-IBV-97006156/2020|EPI_ISL_537303|	····················	····················	····················	····················	NNNNNNNNNNNNNNNNNNNN	NNNNNNNNNNNNNNNNNNNN	1
H	2921 hCoV-19/Spain/CL-IBV-97006157/2020|EPI_ISL_537304|	····················	····················	····················	····················	NNNNNNNNNNNNNNNNNNNN	····················	1
H	2922 hCoV-19/Spain/CL-IBV-97006158/2020|EPI_ISL_537305|	····················	····················	····················	····················	NNNNNNNNNNNNNNNNNNNN	NNNNNNNNNNNNNNNNNNNN	1
H	2923 hCoV-19/Spain/CL-IBV-97006159/2020|EPI_ISL_537306|	····················	····················	····················	····················	NNNNNNNNNNNNNNNNNNNN	NNNNNNNNNNNNNNNNNNNN	1
H	2924 hCoV-19/Spain/CL-IBV-97006160/2020|EPI_ISL_537307|	····················	····················	····················	····················	NNNNNNNNNNNNNNNNNNNN	NNNNNNNNNNNNNNNNNNNN	1
H	2925 hCoV-19/Spain/CL-IBV-97006161/2020|EPI_ISL_537308|	····················	····················	····················	····················	NNNNNNNNNNNNNNNNNNNN	NNNNNNNNNNNNNNNNNNNN	1
H	2926 hCoV-19/Spain/CL-IBV-97006162/2020|EPI_ISL_537309|	····················	····················	····················	····················	NNNNNNNNNNNNNNNNNNNN	NNNNNNNNNNNNNNNNNNNN	1
H	2927 hCoV-19/Spain/CL-IBV-97006163/2020|EPI_ISL_537310|	····················	····················	····················	····················	NNNNNNNNNNNNNNNNNNNN	NNNNNNNNNNNNNNNNNNNN	1
H	2928 hCoV-19/Spain/CL-IBV-97006164/2020|EPI_ISL_537311|	····················	····················	····················	····················	NNNNNNNNNNNNNNNNNNNN	NNNNNNNNNNNNNNNNNNNN	1
H	2929 hCoV-19/Spain/CL-IBV-97006165/2020|EPI_ISL_537312|	····················	····················	····················	····················	NNNNNNNNNNNNNNNNNNNN	NNNNNNNNNNNNNNNNNNNN	1
H	2931 hCoV-19/Spain/CL-IBV-97006167/2020|EPI_ISL_537314|	····················	····················	····················	····················	NNNNNNNNNNNNNNNNNNNN	NNNNNNNNNNNNNNNNNNNN	1
H	2932 hCoV-19/Spain/CL-IBV-97006168/2020|EPI_ISL_537315|	····················	····················	····················	····················	NNNNNNNNNNNNNNNNNNNN	NNNNNNNNNNNNNNNNNNNN	1
H	2933 hCoV-19/Spain/CL-IBV-97006169/2020|EPI_ISL_537316|	····················	····················	····················	····················	NNNNNNNNNNNNNNNNNNNN	NNNNNNNNNNNNNNNNNNNN	1
H	2934 hCoV-19/Spain/CL-IBV-97006170/2020|EPI_ISL_537317|	····················	····················	····················	····················	NNNNNNNNNNNNNNNNNNNN	NNNNNNNNNNNNNNNNNNNN	1
H	3034 hCoV-19/USA/TX-HMH-3655/2020|EPI_ISL_543100|2020-0	····················	····················	····················	····················	NNNNNNNNNNNNNNNNNNNN	NNNNNNNNNNNNNNNNNNNN	1
H	3038 hCoV-19/USA/TX-HMH-3662/2020|EPI_ISL_543106|2020-0	····················	····················	····················	····················	NNNNNNNNNNNNNNNNNNNN	NNNNNNNNNNNNNNNNNNNN	1
H	3040 hCoV-19/USA/TX-HMH-3688/2020|EPI_ISL_543108|2020-0	····················	····················	····················	····················	NNNNNNNNNNNNNNNNNNNN	NNNNNNNNNNNNNNNNNNNN	1
H	3041 hCoV-19/USA/TX-HMH-3689/2020|EPI_ISL_543109|2020-0	····················	····················	····················	····················	NNNNNNNNNNNNNNNNNNNN	NNNNNNNNNNNNNNNNNNNN	1
H	3042 hCoV-19/USA/TX-HMH-3690/2020|EPI_ISL_543110|2020-0	····················	····················	····················	····················	NNNNNNNNNNNNNNNNNNNN	NNNNNNNNNNNNNNNNNNNN	1
H	3043 hCoV-19/USA/TX-HMH-3692/2020|EPI_ISL_543111|2020-0	····················	····················	····················	····················	NNNNNNNNNNNNNNNNNNNN	NNNNNNNNNNNNNNNNNNNN	1
H	3044 hCoV-19/USA/TX-HMH-3693/2020|EPI_ISL_543112|2020-0	····················	····················	····················	····················	NNNNNNNNNNNNNNNNNNNN	NNNNNNNNNNNNNNNNNNNN	1
H	3045 hCoV-19/USA/TX-HMH-3695/2020|EPI_ISL_543114|2020-0	····················	····················	····················	····················	NNNNNNNNNNNNNNNNNNNN	NNNNNNNNNNNNNNNNNNNN	1
H	3047 hCoV-19/USA/TX-HMH-3698/2020|EPI_ISL_543117|2020-0	····················	····················	····················	····················	·············N······	NN·····NN···········	1
H	3048 hCoV-19/USA/TX-HMH-3700/2020|EPI_ISL_543118|2020-0	····················	····················	····················	····················	NNNNNNNNNNNNNNNNNNNN	NNNNNNNNNNNNNNNNNNNN	1
H	3049 hCoV-19/USA/TX-HMH-3701/2020|EPI_ISL_543119|2020-0	····················	····················	····················	····················	NNNNNNNNNNNNNNNNNNNN	NNNNNNNNNNNNNNNNNNNN	1
H	3050 hCoV-19/USA/TX-HMH-3702/2020|EPI_ISL_543120|2020-0	····················	····················	····················	····················	NNNNNNNNNNNNNNNNNNNN	NNNNNNNNNNNNNNNNNNNN	1
H	3051 hCoV-19/USA/TX-HMH-3704/2020|EPI_ISL_543122|2020-0	····················	····················	····················	····················	NNNNNNNNNNNNNNNNNNNN	NNNNNNNNNNNNNNNNNNNN	1
H	3053 hCoV-19/USA/TX-HMH-3707/2020|EPI_ISL_543124|2020-0	····················	····················	····················	····················	NNNNNNNNNNNNNNNNNNNN	NNNNNNNNNNNNNNNNNNNN	1
H	3055 hCoV-19/USA/TX-HMH-3713/2020|EPI_ISL_543130|2020-0	····················	····················	····················	····················	······N····NN·NNNN··	NN·N···NN·NN······N·	1
H	3059 hCoV-19/USA/TX-HMH-3718/2020|EPI_ISL_543135|2020-0	····················	·····A··············	····················	····················	NNNNNNNNNNNNNNNNNNNN	NNNNNNNNNNNNNNNNNNNN	1
H	3060 hCoV-19/USA/TX-HMH-3721/2020|EPI_ISL_543137|2020-0	····················	····················	····················	····················	NNNNNNNNNNNNNNNNNNNN	NNNNNNNNNNNNNNNNNNNN	1
H	3064 hCoV-19/USA/TX-HMH-3736/2020|EPI_ISL_543148|2020-0	····················	····················	····················	····················	NNNNNNNNNNNNNNNNNNNN	NNNNNNNNNNNNNNNNNNNN	1
H	3069 hCoV-19/USA/TX-HMH-3791/2020|EPI_ISL_543196|2020-0	····················	····················	····················	····················	NNNNNNNNNNNNNNNNNNNN	NNNNNNNNNNNNNNNNNNNN	1
```

| | | | | | | | | | |
|---|---|---|---|---|---|---|---|---|---|
| H | 3071 | hCoV-19/USA/TX-HMH-3793/2020\|EPI_ISL_543198\|2020-0 | ···················· | ···················· | ···················· | ···················· | NNNNNNNNNNNNNNNNNNNN | NNNNNNNNNNNNNNNNNNNN | 1 |
| H | 3193 | hCoV-19/Canada/QC-LSPQ-L00234871/2020\|EPI_ISL_5360 | ···················· | ···················· | ···················· | ···················· | ·NNNNNNNNNNNNNNNNNNN | ···················· | 1 |
| H | 3213 | hCoV-19/Spain/CL-IBV-97006172/2020\|EPI_ISL_537319\| | ···················· | ···················· | ···················· | ···················· | NNNNNNNNNNNNNNNNNNNN | NNNNNNNNNNNNNNNNNNNN | 1 |
| H | 3214 | hCoV-19/Spain/CL-IBV-97006173/2020\|EPI_ISL_537320\| | ···················· | ···················· | ···················· | ···················· | NNNNNNNNNNNNNNNNNNNN | NNNNNNNNNNNNNNNNNNNN | 1 |
| H | 3215 | hCoV-19/Spain/CL-IBV-97006174/2020\|EPI_ISL_537321\| | ···················· | ···················· | ···················· | ···················· | NNNNNNNNNNNNNNNNNNNN | NNNNNNNNNNNNNNNNNNNN | 1 |
| H | 3216 | hCoV-19/Spain/CL-IBV-97006175/2020\|EPI_ISL_537322\| | ···················· | ···················· | ···················· | ···················· | ·NN················· | NNNNN········NN····· | 1 |
| H | 3217 | hCoV-19/Spain/CL-IBV-97006177/2020\|EPI_ISL_537324\| | ···················· | ···················· | ···················· | ···················· | NNNNNNNNNNNNNNNNNNNN | ···················· | 1 |
| H | 3218 | hCoV-19/Spain/CL-IBV-97006179/2020\|EPI_ISL_537326\| | ···················· | ···················· | ···················· | ···················· | NNNNNNNNNNNNNNNNNNNN | NNNNNNNNNNNNNNNNNNNN | 1 |
| H | 3219 | hCoV-19/Spain/CL-IBV-97006180/2020\|EPI_ISL_537327\| | ···················· | ···················· | ···················· | ···················· | NNNNNNNNNNNNNNNNNNNN | NNNNNNNNNNNNNNNNNNNN | 1 |
| H | 3220 | hCoV-19/Spain/CL-IBV-97006184/2020\|EPI_ISL_537330\| | ···················· | ···················· | ···················· | ···················· | NNNNNNNNNNNNNNNNNNNN | NNNNNNNNNNNNNNNNNNNN | 1 |
| H | 3221 | hCoV-19/Spain/CL-IBV-97006185/2020\|EPI_ISL_537331\| | ···················· | ···················· | ···················· | ···················· | NNNNNNNNNNNNNNNNNNNN | NNNNNNNNNNNNNNNNNNNN | 1 |
| H | 3222 | hCoV-19/Spain/CL-IBV-97006186/2020\|EPI_ISL_537332\| | ···················· | ···················· | ···················· | ···················· | NNNNNNNNNNNNNNNNNNNN | NNNNNNNNNNNNNNNNNNNN | 1 |
| H | 3223 | hCoV-19/Spain/CL-IBV-97006188/2020\|EPI_ISL_537334\| | ···················· | ···················· | ···················· | ···················· | NNNNNNNNNNNNNNNNNNNN | NNNNNNNNNNNNNNNNNNNN | 1 |
| H | 3224 | hCoV-19/Spain/CL-IBV-97006189/2020\|EPI_ISL_537335\| | ···················· | ···················· | ···················· | ···················· | NNNNNNNNNNNNNNNNNNNN | NNNNNNNNNNNNNNNNNNNN | 1 |
| H | 3225 | hCoV-19/Spain/CL-IBV-97006190/2020\|EPI_ISL_537336\| | ···················· | ···················· | ···················· | ···················· | NNNNNNNNNNNNNNNNNNNN | NNNNNNNNNNNNNNNNNNNN | 1 |
| H | 3226 | hCoV-19/Spain/CL-IBV-97006191/2020\|EPI_ISL_537337\| | ···················· | ···················· | ···················· | ···················· | NNNNNNNNNNNNNNNNNNNN | NNNNNNNNNNNNNNNNNNNN | 1 |
| H | 3227 | hCoV-19/Spain/CL-IBV-97006192/2020\|EPI_ISL_537338\| | ···················· | ···················· | ···················· | ···················· | NNNNNNNNNNNNNNNNNNNN | NNNNNNNNNNNNNNN····· | 1 |
| H | 3228 | hCoV-19/Spain/CL-IBV-97006193/2020\|EPI_ISL_537339\| | ···················· | ···················· | ···················· | ···················· | NNNNNNNNNNNNNNNNNNNN | NNNNNNNNNNNNNNNNNNNN | 1 |
| H | 3229 | hCoV-19/Spain/CL-IBV-97006194/2020\|EPI_ISL_537340\| | ···················· | ···················· | ···················· | ···················· | NNNNNNNNNNNNNNNNNNNN | NNNNNNNNNNNNNNNNNNNN | 1 |
| H | 3230 | hCoV-19/Spain/CL-IBV-97006195/2020\|EPI_ISL_537341\| | ···················· | ···················· | ···················· | ···················· | NNNNNNNNNNNNNNNNNNNN | NNNNNNNNNNNNNNNNNNNN | 1 |
| H | 3231 | hCoV-19/Spain/CL-IBV-97006196/2020\|EPI_ISL_537342\| | ···················· | ···················· | ···················· | ···················· | NNNNNNNNNNN········· | ···················· | 1 |
| H | 3232 | hCoV-19/Spain/CL-IBV-97006197/2020\|EPI_ISL_537343\| | ···················· | ···················· | ···················· | ···················· | NNNNNNNNNNNNNNNNNNNN | NNNNNNNNNNNNNNNNNNNN | 1 |
| H | 3233 | hCoV-19/Spain/CL-IBV-97006198/2020\|EPI_ISL_537344\| | ···················· | ···················· | ···················· | ···················· | NNNNNNNNNNNNNNNNNNNN | NNNNNNNNNNNNNNNNNNNN | 1 |
| H | 3234 | hCoV-19/Spain/CL-IBV-97006200/2020\|EPI_ISL_537346\| | ···················· | ···················· | ···················· | ···················· | NNNNNNNNNNNNNNNNNNNN | NNNNNNNNNNNNNNNNNNNN | 1 |
| H | 3235 | hCoV-19/Spain/CL-IBV-97006201/2020\|EPI_ISL_537347\| | ···················· | ···················· | ···················· | ···················· | NNNNNNN······NNNNNNN | NN·NNNN············· | 1 |
| H | 3236 | hCoV-19/Spain/CL-IBV-97006202/2020\|EPI_ISL_537348\| | ···················· | ···················· | ···················· | ···················· | NNNNNNNNNNNNNNNNNNNN | NNNNNNNNNNNNNNNNNNNN | 1 |
| H | 3237 | hCoV-19/Spain/CL-IBV-97006203/2020\|EPI_ISL_537349\| | ···················· | ···················· | ···················· | ···················· | NNNNNNNNNNNNNNNNNNNN | NNNNNNNNNNNNNNNNNNNN | 1 |
| H | 3238 | hCoV-19/Spain/CL-IBV-97006204/2020\|EPI_ISL_537350\| | ···················· | ···················· | ···················· | ···················· | NNNNNNNNNNNNNNNNNNNN | NNNNNNNNNNNNNNNNNNNN | 1 |
| H | 3239 | hCoV-19/Spain/CL-IBV-97006205/2020\|EPI_ISL_537351\| | ···················· | ···················· | ···················· | ···················· | NNNNNNNNNNNNNNNNNNNN | NNNNNNNNNNNNNNNNNNNN | 1 |
| H | 3240 | hCoV-19/Spain/CL-IBV-97006206/2020\|EPI_ISL_537352\| | ···················· | ···················· | ···················· | ···················· | NNNNNNNNNNNNNNNNNNNN | NNNNNNNNNNNNNNNNNNNN | 1 |
| H | 3241 | hCoV-19/Spain/CL-IBV-97006207/2020\|EPI_ISL_537353\| | ···················· | ···················· | ···················· | ···················· | NNNNNNNNNNNNNNNNNNNN | NNNNNNNNNNNNNNNNNNNN | 1 |
| H | 3242 | hCoV-19/Spain/CL-IBV-97006208/2020\|EPI_ISL_537354\| | ···················· | ···················· | ···················· | ···················· | NNNNNNNNNNNNNNNNNNNN | NNNNNNNNNNNNNNNNNNNN | 1 |
| H | 3243 | hCoV-19/Spain/CL-IBV-97006209/2020\|EPI_ISL_537355\| | ···················· | ···················· | ···················· | ···················· | NNNNNNNNNNNNNNNNNNNN | NNNNNNNNNNNNNNNNNNNN | 1 |
| H | 3244 | hCoV-19/Spain/CL-IBV-97006211/2020\|EPI_ISL_537357\| | ···················· | ···················· | ···················· | ···················· | NNNNNNNNNNNNNNNNNNNN | NNNNNNNNNNNNNNNNNNNN | 1 |
| H | 3245 | hCoV-19/Spain/CL-IBV-97006212/2020\|EPI_ISL_537358\| | ···················· | ···················· | ···················· | ···················· | NNNNNNNNNNNNNNNNNNNN | ·NNNNNNNNNNNNNNNNNNN | 1 |
| H | 3246 | hCoV-19/Spain/CL-IBV-97006213/2020\|EPI_ISL_537359\| | ···················· | ···················· | ···················· | ···················· | ··NNNNNNNNNNNNNNNNNN | ···················· | 1 |
| H | 3247 | hCoV-19/Spain/CL-IBV-97006214/2020\|EPI_ISL_537360\| | ···················· | ···················· | ···················· | ···················· | ··NNNNN············· | ···················· | 1 |
| H | 3248 | hCoV-19/Spain/CL-IBV-97006215/2020\|EPI_ISL_537361\| | ···················· | ···················· | ···················· | ···················· | NNNNNNNNNNNNNNNNNNNN | NNNNNNNNNNNNNNNNNNNN | 1 |
| H | 3249 | hCoV-19/Spain/CL-IBV-97006216/2020\|EPI_ISL_537362\| | ···················· | ···················· | ···················· | ···················· | ·NNNNNN············· | NNNNNNNNNNNNNNN··NNN | 1 |
| H | 3251 | hCoV-19/Spain/CL-IBV-97006219/2020\|EPI_ISL_537365\| | ···················· | ···················· | ···················· | ···················· | NNNNNNNNNNNNNNNNNNNN | NNNNNNNNNNNNNNNNNNNN | 1 |
| H | 3252 | hCoV-19/Spain/CL-IBV-97006220/2020\|EPI_ISL_537366\| | ···················· | ···················· | ···················· | ···················· | NNNNNNNNNNNNNNNNNNNN | NNNNNNNNNNNNNNNNNNNN | 1 |
| H | 3253 | hCoV-19/Spain/CL-IBV-97006221/2020\|EPI_ISL_537367\| | ···················· | ···················· | ···················· | ···················· | NNNNNNNNNNNN········ | NNNNNNNNNNNNNNN····· | 1 |
| H | 3254 | hCoV-19/Spain/CL-IBV-97006222/2020\|EPI_ISL_537368\| | ···················· | ···················· | ···················· | ···················· | NNNNNNNNNNNNNNNNNNNN | N··················· | 1 |
| H | 3255 | hCoV-19/Spain/CL-IBV-97006223/2020\|EPI_ISL_537369\| | ···················· | ···················· | ···················· | ···················· | NNNNNNNNNNNNNNNNNNNN | NNNNNNNNNNNNNNNNNNNN | 1 |
| H | 3256 | hCoV-19/Spain/CL-IBV-97006226/2020\|EPI_ISL_537372\| | ···················· | ···················· | ···················· | ···················· | NNNNNNNNNNNNNNNNNNNN | NNNNNNNNNNNNNNNNNNNN | 1 |
| H | 3315 | hCoV-19/USA/TX-HMH-3728/2020\|EPI_ISL_543140\|2020-0 | ···················· | ···················· | ···················· | ···················· | NNNNNNNNNNNNNNNNNNNN | NNNNNNNNNNNNNNNNNNNN | 1 |
| H | 3316 | hCoV-19/USA/TX-HMH-3729/2020\|EPI_ISL_543141\|2020-0 | ···················· | ···················· | ···················· | ···················· | NNNNNNNNNNNNNNNNNNNN | NNNNNNNNNNNNNNNNNNNN | 1 |
| H | 3318 | hCoV-19/USA/TX-HMH-3731/2020\|EPI_ISL_543143\|2020-0 | ···················· | ···················· | ···················· | ···················· | NNNNNNNNNNNNNNNNNNNN | NNNNNNNNNNNNNNNNNNNN | 1 |
| H | 3322 | hCoV-19/USA/TX-HMH-3738/2020\|EPI_ISL_543150\|2020-0 | ···················· | ···················· | ···················· | ···················· | ··········N···NNNN·N | NN····NN·NN········· | 1 |
| H | 3324 | hCoV-19/USA/TX-HMH-3743/2020\|EPI_ISL_543155\|2020-0 | ···················· | ·········A·········· | ···················· | ···················· | NNNNNNNNNNNNNNNNNNNN | NNNNNNNNNNNNNNNNNNNN | 1 |
| H | 3325 | hCoV-19/USA/TX-HMH-3744/2020\|EPI_ISL_543156\|2020-0 | ···················· | ···················· | ···················· | ···················· | NNNNNNNNNNNNNNNNNNNN | NNNNNNNNNNNNNNNNNNNN | 1 |
| H | 3327 | hCoV-19/USA/TX-HMH-3746/2020\|EPI_ISL_543158\|2020-0 | ···················· | ···················· | ···················· | ···················· | NNNNNNNNNNNNNNNNNNNN | NNNNNNNNNNNNNNNNNNNN | 1 |
| H | 3411 | hCoV-19/Canada/QC-LSPQ-L00234397/2020\|EPI_ISL_5360 | ···················· | ···················· | ···················· | ···················· | ···················· | NNNNNNNNNNNNNNN····· | 1 |
| H | 3428 | hCoV-19/Spain/CL-IBV-97006224/2020\|EPI_ISL_537370\| | ···················· | ···················· | ···················· | ···················· | NNNNNNNNNNNNNNNNNNNN | NNNNNNNNNNNNNNNNNNNN | 1 |
| H | 3429 | hCoV-19/Spain/CL-IBV-97006225/2020\|EPI_ISL_537371\| | ···················· | ···················· | ···················· | ···················· | NNNNNNNNNNNNNNNNNNNN | NNNNNNNNNNNNNNNNNNNN | 1 |
| H | 3430 | hCoV-19/Spain/CL-IBV-97006227/2020\|EPI_ISL_537373\| | ···················· | ···················· | ···················· | ···················· | NNNNNNNNNNNNNNNNNNNN | NNNNNNNNNNNNNNNNNNNN | 1 |
| H | 3432 | hCoV-19/Spain/CL-IBV-97006231/2020\|EPI_ISL_537377\| | ···················· | ···················· | ···················· | ···················· | NNNNNNNNNNNNNNNNNNNN | NNNNNNNNNNNNNNNNNNNN | 1 |
| H | 3434 | hCoV-19/Spain/GA-IBV-97006347/2020\|EPI_ISL_537381\| | ···················· | ···················· | ···················· | ···················· | NNNNNNNNNNNNNNNNNNNN | NNNNNNNNNNNNNNNNNNNN | 1 |
| H | 3436 | hCoV-19/Spain/RI-IBV-99005841/2020\|EPI_ISL_537384\| | ···················· | ···················· | ···················· | ···················· | NNNNNNNNNNNNNNNNNNNN | NNNNNNNNNNNNNNN····· | 1 |
| H | 3440 | hCoV-19/Spain/RI-IBV-99005851/2020\|EPI_ISL_537394\| | ···················· | ···················· | ···················· | ···················· | NNNNNNNNNNNNNNNNNNNN | NNNNNNNNNNNNNNNNN··· | 1 |
| H | 3441 | hCoV-19/Spain/RI-IBV-99005852/2020\|EPI_ISL_537395\| | ···················· | ···················· | ···················· | ···················· | NNNNNNNNNNNNNNNNNNNN | NNNNNNNNNNNNNNNNNNNN | 1 |
| H | 3442 | hCoV-19/Spain/RI-IBV-99005854/2020\|EPI_ISL_537397\| | ···················· | ···················· | ···················· | ···················· | ··NNNNN······NNNN··· | ···················· | 1 |
| H | 3443 | hCoV-19/Spain/RI-IBV-99005855/2020\|EPI_ISL_537398\| | ···················· | ···················· | ···················· | ···················· | NNNNNNNNNNNNNNNNNNNN | NNNNNNNNNNNNNNNNNNNN | 1 |
| H | 3444 | hCoV-19/Spain/RI-IBV-99005856/2020\|EPI_ISL_537399\| | ···················· | ···················· | ···················· | ···················· | NNNNNNNNNNNNNNNNNNNN | NNNNNNNNNNNNNNN····N | 1 |
| H | 3535 | hCoV-19/USA/TX-HMH-3581/2020\|EPI_ISL_543039\|2020-0 | ···················· | ···················· | ···················· | ···················· | NNNNNNNNNNNNNNNNNNNN | NNNNNNNNNNNNNNNNNNNN | 1 |
| H | 3537 | hCoV-19/USA/TX-HMH-3584/2020\|EPI_ISL_543041\|2020-0 | ···················· | ···················· | ···················· | ···················· | NNNNNNNNNNNNNNNNNNNN | NNNNNNNNNNNNNNNNNNNN | 1 |
| H | 3538 | hCoV-19/USA/TX-HMH-3585/2020\|EPI_ISL_543042\|2020-0 | ···················· | ···················· | ···················· | ···················· | NNNNNNNNNNNNNNNNNNNN | NNNNNNNNNNNNNNNNNNNN | 1 |
| H | 3539 | hCoV-19/USA/TX-HMH-3586/2020\|EPI_ISL_543043\|2020-0 | ···················· | ···················· | ···················· | ···················· | ·······N······NN···· | NN····NN············ | 1 |
| H | 3540 | hCoV-19/USA/TX-HMH-3587/2020\|EPI_ISL_543044\|2020-0 | ···················· | ···················· | ···················· | ···················· | NNNNNNNNNNNNNNNNNNNN | NNNNNNNNNNNNNNNNNNNN | 1 |
| H | 3541 | hCoV-19/USA/TX-HMH-3588/2020\|EPI_ISL_543045\|2020-0 | ···················· | ···················· | ···················· | ···················· | NNNNNNNNNNNNNNNNNNNN | NNNNNNNNNNNNNNNNNNNN | 1 |
| H | 3542 | hCoV-19/USA/TX-HMH-3590/2020\|EPI_ISL_543047\|2020-0 | ···················· | ·············A······ | ···················· | ···················· | NNNNNNNNNNNNNNNNNNNN | NNNNNNNNNNNNNNNNNNNN | 1 |
| H | 3543 | hCoV-19/USA/TX-HMH-3593/2020\|EPI_ISL_543049\|2020-0 | ···················· | ···················· | ···················· | ···················· | NNNNNNNNNNNNNNNNNNNN | NNNNNNNNNNNNNNNNNNNN | 1 |
| H | 3545 | hCoV-19/USA/TX-HMH-3596/2020\|EPI_ISL_543052\|2020-0 | ···················· | ···················· | ···················· | ···················· | ··············N····· | NNNN···NNNNNN·····NN | 1 |
| H | 3546 | hCoV-19/USA/TX-HMH-3597/2020\|EPI_ISL_543053\|2020-0 | ···················· | ···················· | ···················· | ···················· | NNNNNNNNNNNNNNNNNNNN | NNNNNNNNNNNNNNNNNNNN | 1 |
| H | 3547 | hCoV-19/USA/TX-HMH-3598/2020\|EPI_ISL_543054\|2020-0 | ···················· | ···················· | ···················· | ···················· | NNNNNNNNNNNNNNNNNNNN | NNNNNNNNNNNNNNNNNNNN | 1 |
| H | 3549 | hCoV-19/USA/TX-HMH-3600/2020\|EPI_ISL_543056\|2020-0 | ···················· | ···················· | ···················· | ···················· | NNNNNNNNNNNNNNNNNNNN | NNNNNNNNNNNNNNNNNNNN | 1 |
| H | 3550 | hCoV-19/USA/TX-HMH-3601/2020\|EPI_ISL_543057\|2020-0 | ···················· | ···················· | ···················· | ···················· | NNNNNNNNNNNNNNNNNNNN | NNNNNNNNNNNNNNNNNNNN | 1 |
| H | 3551 | hCoV-19/USA/TX-HMH-3605/2020\|EPI_ISL_543060\|2020-0 | ···················· | ···················· | ···················· | ···················· | NNNNNNNNNNNNNNNNNNNN | NNNNNNNNNNNNNNNNNNNN | 1 |
| H | 3552 | hCoV-19/USA/TX-HMH-3606/2020\|EPI_ISL_543061\|2020-0 | ···················· | ···················· | ···················· | ···················· | NNNNNNNNNNNNNNNNNNNN | NNNNNNNNNNNNNNNNNNNN | 1 |
| H | 3557 | hCoV-19/USA/TX-HMH-3612/2020\|EPI_ISL_543066\|2020-0 | ···················· | ······A············· | ···················· | ···················· | NNNNNNNNNNNNNNNNNNNN | NNNNNNNNNNNNNNNNNNNN | 1 |

| | | | | | | | | | |
|---|---|---|---|---|---|---|---|---|---|
| H | 3558 | hCoV-19/USA/TX-HMH-3613/2020\|EPI_ISL_543067\|2020-0 | ···················· | ···················· | ···················· | ···················· | NNNNNNNNNNNNNNNNNNNN | NNNNNNNNNNNNNNNNNNNN | 1 |
| H | 3559 | hCoV-19/USA/TX-HMH-3614/2020\|EPI_ISL_543068\|2020-0 | ···················· | ···················· | ···················· | ···················· | NNNNNNNNNNNNNNNNNNNN | NNNNNNNNNNNNNNNNNNNN | 1 |
| H | 3564 | hCoV-19/USA/TX-HMH-3621/2020\|EPI_ISL_543074\|2020-0 | ···················· | ···················· | ···················· | ···················· | NNNNNNNNNNNNNNNNNNNN | NNNNNNNNNNNNNNNNNNNN | 1 |
| H | 3569 | hCoV-19/USA/TX-HMH-3629/2020\|EPI_ISL_543081\|2020-0 | ···················· | ···················· | ···················· | ···················· | NNNNNNNNNNNNNNNNNNNN | NNNNNNNNNNNNNNNNNNNN | 1 |
| H | 3570 | hCoV-19/USA/TX-HMH-3632/2020\|EPI_ISL_543082\|2020-0 | ···················· | ···················· | ···················· | ···················· | NNNNNNNNNNNNNNNNNNNN | NNNNNNNNNNNNNNNNNNNN | 1 |
| H | 3571 | hCoV-19/USA/TX-HMH-3634/2020\|EPI_ISL_543084\|2020-0 | ···················· | ···················· | ···················· | ···················· | NNNNNNNNNNNNNNNNNNNN | NNNNNNNNNNNNNNNNNNNN | 1 |
| H | 3573 | hCoV-19/USA/TX-HMH-3638/2020\|EPI_ISL_543086\|2020-0 | ···················· | ···················· | ···················· | ···················· | ··············N····· | NN·····NN··········· | 1 |
| H | 3576 | hCoV-19/USA/TX-HMH-3641/2020\|EPI_ISL_543089\|2020-0 | ···················· | ···················· | ···················· | ···················· | NNNNNNNNNNNNNNNNNNNN | NNNNNNNNNNNNNNNNNNNN | 1 |
| H | 3581 | hCoV-19/USA/TX-HMH-3648/2020\|EPI_ISL_543094\|2020-0 | ···················· | ···················· | ···················· | ···················· | NNNNNNNNNNNNNNNNNNNN | NNNNNNNNNNNNNNNNNNNN | 1 |
| H | 3582 | hCoV-19/USA/TX-HMH-3649/2020\|EPI_ISL_543095\|2020-0 | ···················· | ···················· | ···················· | ···················· | NNNNNNNNNNNNNNNNNNNN | NNNNNNNNNNNNNNNNNNNN | 1 |
| H | 3583 | hCoV-19/USA/TX-HMH-3651/2020\|EPI_ISL_543097\|2020-0 | ···················· | ···················· | ···················· | ···················· | NNNNNNNNNNNNNNNNNNNN | NNNNNNNNNNNNNNNNNNNN | 1 |
| H | 3762 | hCoV-19/Spain/CL-IBV-97006140/2020\|EPI_ISL_537288\| | ···················· | ···················· | ···················· | ···················· | NNNNNNNNNNNNNNNNNNNN | NNNNNNNNNNNNNNNNNNNN | 1 |
| H | 3763 | hCoV-19/Spain/CL-IBV-97006141/2020\|EPI_ISL_537289\| | ···················· | ···················· | ···················· | ···················· | NNNNNNNNNNNNNNNNNNNN | NNNNNNNNNNNNNNNNNNNN | 1 |
| H | 3764 | hCoV-19/Spain/CL-IBV-97006142/2020\|EPI_ISL_537290\| | ···················· | ···················· | ···················· | ···················· | NNNNNNNNNNNNNNNNNNNN | NNNNNNNNNNNNNNNNNNNN | 1 |
| H | 3765 | hCoV-19/Spain/CL-IBV-97006144/2020\|EPI_ISL_537291\| | ···················· | ···················· | ···················· | ···················· | NNNNNNNNNNNNNNNNNNNN | NNNNNNNNNNNNNNNNNNNN | 1 |
| H | 3766 | hCoV-19/Spain/CL-IBV-97006146/2020\|EPI_ISL_537293\| | ···················· | ···················· | ···················· | ···················· | NNNNNNNNNNNNNNNN···· | ···················· | 1 |
| H | 3767 | hCoV-19/Spain/CL-IBV-97006147/2020\|EPI_ISL_537294\| | ···················· | ···················· | ···················· | ···················· | NNNNNNNNNNNNNNNNNNNN | ·····NNNNNNNNNN····· | 1 |
| H | 3768 | hCoV-19/Spain/CL-IBV-97006148/2020\|EPI_ISL_537295\| | ···················· | ···················· | ···················· | ···················· | NNNNNNNNNNNNNNNNNNNN | NNNNNNNNNNNNNNNNNNNN | 1 |
| H | 3770 | hCoV-19/Spain/CL-IBV-97006150/2020\|EPI_ISL_537297\| | ···················· | ···················· | ···················· | ···················· | NNNNNNNNNNNNNNNNNNNN | ···················· | 1 |
| H | 3771 | hCoV-19/Spain/CL-IBV-97006151/2020\|EPI_ISL_537298\| | ···················· | ···················· | ···················· | ···················· | NNNNNNNNNNNNNNNNNNNN | NNNNNNNNNNNNNNNNNNNN | 1 |
| H | 3772 | hCoV-19/Spain/CL-IBV-97006152/2020\|EPI_ISL_537299\| | ···················· | ···················· | ···················· | ···················· | NNNNNNNNNNNNNNNNNNNN | NNNNNNNNNNNNNNNNNNNN | 1 |
| H | 3818 | hCoV-19/USA/TX-HMH-3545/2020\|EPI_ISL_543008\|2020-0 | ···················· | ···················· | ···················· | ···················· | NNNNNNNNNNNNNNNNNNNN | NNNNNNNNNNNNNNNNNNNN | 1 |
| H | 3820 | hCoV-19/USA/TX-HMH-3549/2020\|EPI_ISL_543011\|2020-0 | ···················· | ···················· | ···················· | ···················· | NNNNNNNNNNNNNNNNNNNN | NNNNNNNNNNNNNNNNNNNN | 1 |
| H | 3821 | hCoV-19/USA/TX-HMH-3550/2020\|EPI_ISL_543012\|2020-0 | ···················· | ···················· | ···················· | ···················· | NNNNNNNNNNNNNNNNNNNN | NNNNNNNNNNNNNNNNNNNN | 1 |
| H | 3822 | hCoV-19/USA/TX-HMH-3552/2020\|EPI_ISL_543014\|2020-0 | ···················· | ···················· | ···················· | ···················· | NNNNNNNNNNNNNNNNNNNN | NNNNNNNNNNNNNNNNNNNN | 1 |
| H | 3826 | hCoV-19/USA/TX-HMH-3557/2020\|EPI_ISL_543019\|2020-0 | ···················· | ···················· | ···················· | ···················· | NNNNNNNNNNNNNNNNNNNN | NNNNNNNNNNNNNNNNNNNN | 1 |
| H | 3827 | hCoV-19/USA/TX-HMH-3558/2020\|EPI_ISL_543020\|2020-0 | ···················· | ···················· | ···················· | ···················· | NNNNNNNNNNNNNNNNNNNN | NNNNNNNNNNNNNNNNNNNN | 1 |
| H | 3829 | hCoV-19/USA/TX-HMH-3562/2020\|EPI_ISL_543024\|2020-0 | ···················· | ···················· | ···················· | ···················· | NNNNNNNNNNNNNNNNNNNN | NNNNNNNNNNNNNNNNNNNN | 1 |
| H | 3830 | hCoV-19/USA/TX-HMH-3563/2020\|EPI_ISL_543025\|2020-0 | ···················· | ···················· | ···················· | ···················· | NNNNNNNNNNNNNNNNNNNN | NNNNNNNNNNNNNNNNNNNN | 1 |
| H | 3831 | hCoV-19/USA/TX-HMH-3566/2020\|EPI_ISL_543027\|2020-0 | ···················· | ·····A·············· | ···················· | ···················· | NNNNNNNNNNNNNNNNNNNN | NNNNNNNNNNNNNNNNNNNN | 1 |
| H | 3832 | hCoV-19/USA/TX-HMH-3569/2020\|EPI_ISL_543029\|2020-0 | ···················· | ···················· | ···················· | ···················· | NNNNNNNNNNNNNNNNNNNN | NNNNNNNNNNNNNNNNNNNN | 1 |
| H | 3835 | hCoV-19/USA/TX-HMH-3573/2020\|EPI_ISL_543033\|2020-0 | ···················· | ···················· | ···················· | ···················· | NNNNNNNNNNNNNNNNNNNN | NNNNNNNNNNNNNNNNNNNN | 1 |
| H | 3836 | hCoV-19/USA/TX-HMH-3574/2020\|EPI_ISL_543034\|2020-0 | ···················· | ···················· | ···················· | ···················· | NNNNNNNNNNNNNNNNNNNN | NNNNNNNNNNNNNNNNNNNN | 1 |
| H | 3837 | hCoV-19/USA/TX-HMH-3575/2020\|EPI_ISL_543035\|2020-0 | ···················· | ···················· | ···················· | ···················· | NNNNNNNNNNNNNNNNNNNN | NNNNNNNNNNNNNNNNNNNN | 1 |
| H | 3839 | hCoV-19/USA/TX-HMH-3578/2020\|EPI_ISL_543037\|2020-0 | ···················· | ···················· | ···················· | ···················· | NNNNNNNNNNNNNNNNNNNN | NNNNNNNNNNNNNNNNNNNN | 1 |
| H | 4385 | hCoV-19/Wales/PHWC-16A21C/2020\|EPI_ISL_535217\|2020 | ···················· | ···················· | ···················· | ···················· | ········NNNNNNNNN··· | ···················· | 1 |
| H | 4389 | hCoV-19/Wales/PHWC-16A294/2020\|EPI_ISL_535225\|2020 | ···················· | ···················· | ···················· | ···················· | NNNNNNNNNNNNNNNNNNNN | ···················· | 1 |
| H | 4391 | hCoV-19/Wales/PHWC-16A2B2/2020\|EPI_ISL_535227\|2020 | ···················· | ···················· | ···················· | ···················· | NNNNNNNNNNNNNNNNNNNN | ···················· | 1 |
| H | 4393 | hCoV-19/Wales/PHWC-16A2EF/2020\|EPI_ISL_535230\|2020 | ···················· | ···················· | ···················· | ···················· | ·NNNNNNNNNNNNNNNNNNN | NNNNNNNNNNNNNNNNNNNN | 1 |
| H | 4395 | hCoV-19/Wales/PHWC-16A328/2020\|EPI_ISL_535234\|2020 | ···················· | ···················· | ···················· | ···················· | NNNNNNNNNNNNNNNNNNNN | ···················· | 1 |
| H | 4397 | hCoV-19/Wales/PHWC-16A364/2020\|EPI_ISL_535238\|2020 | ···················· | ···················· | ···················· | ···················· | NNNNNNNNNNNNNNNNNNNN | ···················· | 1 |
| H | 4399 | hCoV-19/Wales/PHWC-16A3A0/2020\|EPI_ISL_535242\|2020 | ···················· | ···················· | ···················· | ···················· | NNNNNNNNNNNNNNNNNNNN | NNNNNNNNNNNNNNNNNNNN | 1 |
| H | 4401 | hCoV-19/Wales/PHWC-16A3FB/2020\|EPI_ISL_535246\|2020 | ···················· | ···················· | ···················· | ···················· | NNNNNNNNNNNNNNNNNNNN | ···················· | 1 |
| H | 4402 | hCoV-19/Wales/PHWC-16A407/2020\|EPI_ISL_535247\|2020 | ···················· | ···················· | ···················· | ···················· | NNNNNNNNNNNNNNNNNNNN | ···················· | 1 |
| H | 4403 | hCoV-19/Wales/PHWC-16A416/2020\|EPI_ISL_535248\|2020 | ···················· | ···················· | ···················· | ···················· | NNNNN··············· | ···················· | 1 |
| H | 4478 | hCoV-19/USA/WI-UW-1216/2020\|EPI_ISL_536589\|2020-08 | ···················· | NNNNNNNNNNNNNNNNNNNN | ···················· | ···················· | ···················· | ···················· | 1 |
| H | 4494 | hCoV-19/Spain/RI-IBV-98002306/2020\|EPI_ISL_537829\| | ···················· | ···················· | ···················· | ···················· | NNNNNNNNNNNNNNNNNNNN | ···················· | 1 |
| H | 4495 | hCoV-19/Spain/RI-IBV-98002308/2020\|EPI_ISL_537831\| | ···················· | ···················· | ···················· | ···················· | NNNNNNNNNNNNNNNNNNNN | NNNNNNNNNNNNNNNNNNNN | 1 |
| H | 4498 | hCoV-19/Spain/RI-IBV-98002318/2020\|EPI_ISL_537838\| | ···················· | ···················· | ···················· | ···················· | NNNNNNN············· | NNNNNNNNNNNNNNNNNNNN | 1 |
| H | 4499 | hCoV-19/Spain/RI-IBV-98002320/2020\|EPI_ISL_537839\| | ···················· | ···················· | ···················· | ···················· | NNNNNNNNNNNNNNNNNNNN | NNNNNNNNNNNNNNN·NNNN | 1 |
| H | 4503 | hCoV-19/Spain/RI-IBV-98002325/2020\|EPI_ISL_537844\| | ···················· | ···················· | ···················· | ···················· | NNNNNNN············· | ···················· | 1 |
| H | 4504 | hCoV-19/Spain/RI-IBV-98002328/2020\|EPI_ISL_537845\| | ···················· | ···················· | ···················· | ···················· | NNNNNNNNNNNNNNNNNNNN | NNNNNNNNNNNNNNNNNNNN | 1 |
| H | 4505 | hCoV-19/Spain/RI-IBV-98002334/2020\|EPI_ISL_537847\| | ···················· | ···················· | ···················· | ···················· | NNNNNNNNNNNNNNNNNNNN | NNNNNNNNNNNNNNNNNNNN | 1 |
| H | 4506 | hCoV-19/Spain/RI-IBV-98002336/2020\|EPI_ISL_537849\| | ···················· | ···················· | ···················· | ···················· | NNNNNNNNNNNNNNNNNNNN | NNNNNNNN···NNNNNNNNN | 1 |
| H | 4507 | hCoV-19/Spain/RI-IBV-98002337/2020\|EPI_ISL_537850\| | ···················· | ···················· | ···················· | ···················· | NNNNNNNNNNNNNNNNNNNN | NNNNNNNNNNNNNNNNNNNN | 1 |
| H | 4508 | hCoV-19/Spain/RI-IBV-98002339/2020\|EPI_ISL_537852\| | ···················· | ···················· | ···················· | ···················· | NNNNNNNNNNNNNNNNNNNN | NNNNNNNNNNNNNNNNNNNN | 1 |
| H | 4509 | hCoV-19/Spain/RI-IBV-98002340/2020\|EPI_ISL_537853\| | ···················· | ···················· | ···················· | ···················· | NNNNNNNNNNNNNNNNNNNN | NNNNNNNNNNNNNNNNNNNN | 1 |
| H | 4510 | hCoV-19/Spain/RI-IBV-98002342/2020\|EPI_ISL_537855\| | ···················· | ···················· | ···················· | ···················· | NNNNNNNNNNNNNNNNNNNN | NNNNNNNNNNNNNNN··NNN | 1 |
| H | 4513 | hCoV-19/Spain/RI-IBV-98002354/2020\|EPI_ISL_537858\| | ···················· | ···················· | ···················· | ···················· | NNNNNNNNNNNNNNNNNNNN | NNNNNNNNNNNNNNNNNNNN | 1 |
| H | 4515 | hCoV-19/Spain/RI-IBV-98002367/2020\|EPI_ISL_537863\| | ···················· | ···················· | ···················· | ···················· | NNNNNNNNNNNNNNNNNNNN | NNNNNNNNNNNNNNNNNNNN | 1 |
| H | 4516 | hCoV-19/Spain/RI-IBV-98002368/2020\|EPI_ISL_537864\| | ···················· | ···················· | ···················· | ···················· | NNNNNNNNNNNNN······· | NNN················· | 1 |
| H | 4517 | hCoV-19/Spain/RI-IBV-98002369/2020\|EPI_ISL_537865\| | ···················· | ···················· | ···················· | ···················· | NNNNNNNNNNNNNNNNNNNN | NNNNNNNNNNNNNNNNNNNN | 1 |
| H | 4518 | hCoV-19/Spain/RI-IBV-98002370/2020\|EPI_ISL_537866\| | ···················· | ···················· | ···················· | ···················· | NNNNNNNNNNNNNNNNNNNN | NNNNNNNNNNNNNNN····N | 1 |
| H | 4521 | hCoV-19/Spain/RI-IBV-98002384/2020\|EPI_ISL_537873\| | ···················· | ···················· | ···················· | ···················· | NNNNNNNNNNNNNNNNNNNN | NNNNNNNNNNNNNNNNNNNN | 1 |
| H | 4592 | hCoV-19/USA/TX-HMH-4508/2020\|EPI_ISL_543678\|2020-0 | ···················· | ···················· | ···················· | ···················· | NNNNNNNNNNNNNNNNNNNN | NNNNNNNNNNNNNNNNNNNN | 1 |
| H | 4593 | hCoV-19/USA/TX-HMH-4510/2020\|EPI_ISL_543680\|2020-0 | ···················· | ···················· | ···················· | ···················· | NNNNNNNNNNNNNNNNNNNN | NNNNNNNNNNNNNNNNNNNN | 1 |
| H | 4594 | hCoV-19/USA/TX-HMH-4513/2020\|EPI_ISL_543682\|2020-0 | ···················· | ···················· | ···················· | ···················· | NNNNNNNNNNNNNNNNNNNN | NNNNNNNNNNNNNNNNNNNN | 1 |
| H | 4595 | hCoV-19/USA/TX-HMH-4514/2020\|EPI_ISL_543683\|2020-0 | ···················· | ···················· | ···················· | ···················· | NN·····N·NNNNNNN·N·· | ·NNN···NN··········· | 1 |
| H | 4596 | hCoV-19/USA/TX-HMH-4515/2020\|EPI_ISL_543684\|2020-0 | ···················· | ·············A······ | ···················· | ···················· | NNNNNNNNNNNNNNNNNNNN | NNNNNNNNNNNNNNNNNNNN | 1 |
| H | 4598 | hCoV-19/USA/TX-HMH-4519/2020\|EPI_ISL_543688\|2020-0 | ···················· | ···················· | ···················· | ···················· | NNNNNNNNNNNNNNNNNNNN | NNNNNNNNNNNNNNNNNNNN | 1 |
| H | 4599 | hCoV-19/USA/TX-HMH-4521/2020\|EPI_ISL_543689\|2020-0 | ···················· | ···················· | ···················· | ···················· | NNNNNNNNNNNNNNNNNNNN | NNNNNNNNNNNNNNNNNNNN | 1 |
| H | 4602 | hCoV-19/USA/TX-HMH-4527/2020\|EPI_ISL_543693\|2020-0 | ···················· | ···················· | ···················· | ···················· | NNNNNNNNNNNNNNNNNNNN | NNNNNNNNNNNNNNNNNNNN | 1 |
| H | 4603 | hCoV-19/USA/TX-HMH-4529/2020\|EPI_ISL_543694\|2020-0 | ···················· | ···················· | ···················· | ···················· | NNNNNNNNNNNNNNNNNNNN | NNNNNNNNNNNNNNNNNNNN | 1 |
| H | 4726 | hCoV-19/Wales/PHWC-16A4DA/2020\|EPI_ISL_535260\|2020 | ···················· | ···················· | ···················· | ···················· | NNNNNNNNNNNNNN······ | NNNNNNNNNNNNNNN····· | 1 |
| H | 4751 | hCoV-19/USA/WI-UW-1219/2020\|EPI_ISL_536592\|2020-08 | ···················· | ···················· | ···················· | ···················· | NNNNNNNNNNNNNNNNNNNN | NNNNNNNNNNNNNNNNNNNN | 1 |
| H | 4809 | hCoV-19/USA/TX-HMH-4563/2020\|EPI_ISL_543719\|2020-0 | ···················· | ···················· | ···················· | ···················· | NNNNNNNNNNNNNNNNNNNN | NNNNNNNNNNNNNNNNNNNN | 1 |
| H | 4810 | hCoV-19/USA/TX-HMH-4565/2020\|EPI_ISL_543720\|2020-0 | ···················· | ·············A······ | ···················· | ···················· | NNNNNNNNNNNNNNNNNNNN | NNNNNNNNNNNNNNNNNNNN | 1 |
| H | 4811 | hCoV-19/USA/TX-HMH-4567/2020\|EPI_ISL_543721\|2020-0 | ···················· | ·A·················· | ···················· | ···················· | NNNNNNNNNNNNNNNNNNNN | NNNNNNNNNNNNNNNNNNNN | 1 |
| H | 4812 | hCoV-19/USA/TX-HMH-4568/2020\|EPI_ISL_543722\|2020-0 | ···················· | ···················· | ···················· | ···················· | NNNNNNNNNNNNNNNNNNNN | NNNNNNNNNNNNNNNNNNNN | 1 |
| H | 4816 | hCoV-19/USA/TX-HMH-4573/2020\|EPI_ISL_543726\|2020-0 | ···················· | ···················· | ···················· | ···················· | NNNNNNNNNNNNNNNNNNNN | NNNNNNNNNNNNNNNNNNNN | 1 |

| | | | | | | | | | |
|---|---|---|---|---|---|---|---|---|---|
| H | 4817 | hCoV-19/USA/TX-HMH-4575/2020\|EPI_ISL_543727\|2020-0 | ···················· | ···················· | ···················· | ···················· | NNNNNNNNNNNNNNNNNNNN | NNNNNNNNNNNNNNNNNNNN | 1 |
| H | 4818 | hCoV-19/USA/TX-HMH-4578/2020\|EPI_ISL_543728\|2020-0 | ···················· | ···················· | ···················· | ···················· | NNNNNNNNNNNNNNNNNNNN | NNNNNNNNNNNNNNNNNNNN | 1 |
| H | 4821 | hCoV-19/USA/TX-HMH-4581/2020\|EPI_ISL_543731\|2020-0 | ···················· | ···················· | ···················· | ···················· | NNNNNNNNNNNNNNNNNNNN | NNNNNNNNNNNNNNNNNNNN | 1 |
| H | 4823 | hCoV-19/USA/TX-HMH-4583/2020\|EPI_ISL_543733\|2020-0 | ···················· | ···················· | ···················· | ···················· | NNNNNNNNNNNNNNNNNNNN | NNNNNNNNNNNNNNNNNNNN | 1 |
| H | 4826 | hCoV-19/USA/TX-HMH-4586/2020\|EPI_ISL_543736\|2020-0 | ···················· | ···················· | ···················· | ···················· | NNNNNNNNNNNNNNNNNNNN | NNNNNNNNNNNNNNNNNNNN | 1 |
| H | 4828 | hCoV-19/USA/TX-HMH-4589/2020\|EPI_ISL_543738\|2020-0 | ···················· | ·A·················· | ···················· | ···················· | ···················· | NN·N···NN··········· | 1 |
| H | 4829 | hCoV-19/USA/TX-HMH-4591/2020\|EPI_ISL_543739\|2020-0 | ···················· | ···················· | ···················· | ···················· | NN···········N···N·· | NN·NN···········N··· | 1 |
| H | 4831 | hCoV-19/USA/TX-HMH-4596/2020\|EPI_ISL_543742\|2020-0 | ···················· | ···················· | ···················· | ···················· | ··············NN···· | NN·····NN··········· | 1 |
| H | 4832 | hCoV-19/USA/TX-HMH-4597/2020\|EPI_ISL_543743\|2020-0 | ···················· | ···················· | ···················· | ···················· | NNNNNNNNNNNNNNNNNNNN | NNNNNNNNNNNNNNNNNNNN | 1 |
| H | 4834 | hCoV-19/USA/TX-HMH-4600/2020\|EPI_ISL_543746\|2020-0 | ···················· | ·············A······ | ···················· | ···················· | NNNNNNNNNNNNNNNNNNNN | NNNNNNNNNNNNNNNNNNNN | 1 |
| H | 4835 | hCoV-19/USA/TX-HMH-4602/2020\|EPI_ISL_543748\|2020-0 | ···················· | ···················· | ···················· | ···················· | NNNNNNNNNNNNNNNNNNNN | NNNNNNNNNNNNNNNNNNNN | 1 |
| H | 4836 | hCoV-19/USA/TX-HMH-4604/2020\|EPI_ISL_543750\|2020-0 | ···················· | ···················· | ···················· | ···················· | NNNNNNNNNNNNNNNNNNNN | NNNNNNNNNNNNNNNNNNNN | 1 |
| H | 4837 | hCoV-19/USA/TX-HMH-4605/2020\|EPI_ISL_543751\|2020-0 | ···················· | ···················· | ···················· | ···················· | NNNNNNNNNNNNNNNNNNNN | NNNNNNNNNNNNNNNNNNNN | 1 |
| H | 4841 | hCoV-19/USA/TX-HMH-4617/2020\|EPI_ISL_543758\|2020-0 | ···················· | ···················· | ···················· | ···················· | NNNNNNNNNNNNNNNNNNNN | NNNNNNNNNNNNNNNNNNNN | 1 |
| H | 4844 | hCoV-19/USA/TX-HMH-4621/2020\|EPI_ISL_543761\|2020-0 | ···················· | ···················· | ···················· | ···················· | NNNNNNNNNNNNNNNNNNNN | NNNNNNNNNNNNNNNNNNNN | 1 |
| H | 4847 | hCoV-19/USA/TX-HMH-4626/2020\|EPI_ISL_543764\|2020-0 | ···················· | ···················· | ···················· | ···················· | NNNNNNNNNNNNNNNNNNNN | NNNNNNNNNNNNNNNNNNNN | 1 |
| H | 4848 | hCoV-19/USA/TX-HMH-4628/2020\|EPI_ISL_543765\|2020-0 | ···················· | ·············A······ | ···················· | ···················· | NNNNNNNNNNNNNNNNNNNN | NNNNNNNNNNNNNNNNNNNN | 1 |
| H | 4851 | hCoV-19/USA/TX-HMH-4631/2020\|EPI_ISL_543769\|2020-0 | ···················· | ··········A········· | ···················· | ···················· | NNNNNNNNNNNNNNNNNNNN | NNNNNNNNNNNNNNNNNNNN | 1 |
| H | 4853 | hCoV-19/USA/TX-HMH-4635/2020\|EPI_ISL_543772\|2020-0 | ···················· | ···················· | ···················· | ···················· | NNNNNNNNNNNNNNNNNNNN | NNNNNNNNNNNNNNNNNNNN | 1 |
| H | 4858 | hCoV-19/USA/TX-HMH-4640/2020\|EPI_ISL_543777\|2020-0 | ···················· | ···················· | ···················· | ···················· | NNNNNNNNNNNNNNNNNNNN | NNNNNNNNNNNNNNNNNNNN | 1 |
| H | 4861 | hCoV-19/USA/TX-HMH-4655/2020\|EPI_ISL_543783\|2020-0 | ···················· | ·············A······ | ···················· | ···················· | NNNNNNNNNNNNNNNNNNNN | NNNNNNNNNNNNNNNNNNNN | 1 |
| H | 4862 | hCoV-19/USA/TX-HMH-4659/2020\|EPI_ISL_543785\|2020-0 | ···················· | ···················· | ···················· | ···················· | ···················· | ·N·N···NN··········· | 1 |
| H | 4864 | hCoV-19/USA/TX-HMH-4664/2020\|EPI_ISL_543788\|2020-0 | ···················· | ···················· | ···················· | ···················· | NNNNNNNNNNNNNNNNNNNN | NNNNNNNNNNNNNNNNNNNN | 1 |
| H | 5023 | hCoV-19/Spain/CN-IBV-97006507/2020\|EPI_ISL_537715\| | ···················· | ·A·················· | ···················· | ···················· | NNNNNNNNNNNNNNNNNNNN | NNNNNNNNNNNNNNNNNNNN | 1 |
| H | 5025 | hCoV-19/Spain/CN-IBV-97006510/2020\|EPI_ISL_537718\| | ···················· | ···················· | ···················· | ···················· | NNNNNNNNNNNNNNNNNNNN | NNNNNNNNNNNNNNNNNNNN | 1 |
| H | 5026 | hCoV-19/Spain/CN-IBV-97006514/2020\|EPI_ISL_537720\| | ···················· | ·A·················· | ···················· | ···················· | NNNNNNNNNNNNNNNNNNNN | NNNNNNNNNNNNNNNNNNNN | 1 |
| H | 5075 | hCoV-19/USA/TX-HMH-4357/2020\|EPI_ISL_543556\|2020-0 | ···················· | ······A············· | ···················· | ···················· | NNNNNNNNNNNNNNNNNNNN | NNNNNNNNNNNNNNNNNNNN | 1 |
| H | 5076 | hCoV-19/USA/TX-HMH-4358/2020\|EPI_ISL_543557\|2020-0 | ···················· | NNNNNNNNNNNNNNNNNNNN | ···················· | ···················· | ·····N········N··N·· | NN·····NN··········· | 1 |
| H | 5078 | hCoV-19/USA/TX-HMH-4365/2020\|EPI_ISL_543563\|2020-0 | ···················· | ···················· | ···················· | ···················· | NNNNNNNNNNNNNNNNNNNN | NNNNNNNNNNNNNNNNNNNN | 1 |
| H | 5080 | hCoV-19/USA/TX-HMH-4369/2020\|EPI_ISL_543566\|2020-0 | ···················· | ·············A······ | ···················· | ···················· | NNNNNNNNNNNNNNNNNNNN | NNNNNNNNNNNNNNNNNNNN | 1 |
| H | 5084 | hCoV-19/USA/TX-HMH-4373/2020\|EPI_ISL_543570\|2020-0 | ···················· | ···················· | ···················· | ···················· | NNNNNNNNNNNNNNNNNNNN | NNNNNNNNNNNNNNNNNNNN | 1 |
| H | 5090 | hCoV-19/USA/TX-HMH-4383/2020\|EPI_ISL_543577\|2020-0 | ···················· | ···················· | ···················· | ···················· | NNNNNNNNNNNNNNNNNNNN | NNNNNNNNNNNNNNNNNNNN | 1 |
| H | 5091 | hCoV-19/USA/TX-HMH-4385/2020\|EPI_ISL_543579\|2020-0 | ···················· | ···················· | ···················· | ···················· | NNNNNNNNNNNNNNNNNNNN | NNNNNNNNNNNNNNNNNNNN | 1 |
| H | 5096 | hCoV-19/USA/TX-HMH-4392/2020\|EPI_ISL_543584\|2020-0 | ···················· | ···················· | ···················· | ···················· | NNNNNNNNNNNNNNNNNNNN | NNNNNNNNNNNNNNNNNNNN | 1 |
| H | 5099 | hCoV-19/USA/TX-HMH-4398/2020\|EPI_ISL_543587\|2020-0 | ···················· | ···················· | ···················· | ···················· | NNNNNNNNNNNNNNNNNNNN | NNNNNNNNNNNNNNNNNNNN | 1 |
| H | 5100 | hCoV-19/USA/TX-HMH-4399/2020\|EPI_ISL_543588\|2020-0 | ···················· | ···················· | ···················· | ···················· | NNNNNNNNNNNNNNNNNNNN | NNNNNNNNNNNNNNNNNNNN | 1 |
| H | 5104 | hCoV-19/USA/TX-HMH-4403/2020\|EPI_ISL_543592\|2020-0 | ···················· | ···················· | ···················· | ···················· | NNNNNNNNNNNNNNNNNNNN | NNNNNNNNNNNNNNNNNNNN | 1 |
| H | 5105 | hCoV-19/USA/TX-HMH-4404/2020\|EPI_ISL_543593\|2020-0 | ···················· | ···················· | ···················· | ···················· | NNNNNNNNNNNNNNNNNNNN | NNNNNNNNNNNNNNNNNNNN | 1 |
| H | 5107 | hCoV-19/USA/TX-HMH-4407/2020\|EPI_ISL_543596\|2020-0 | ···················· | ···················· | ···················· | ···················· | NNNNNNNNNNNNNNNNNNNN | NNNNNNNNNNNNNNNNNNNN | 1 |
| H | 5113 | hCoV-19/USA/TX-HMH-4539/2020\|EPI_ISL_543702\|2020-0 | ···················· | ···················· | ···················· | ···················· | NNNNNNNNNNNNNNNNNNNN | NNNNNNNNNNNNNNNNNNNN | 1 |
| H | 5116 | hCoV-19/USA/TX-HMH-4541/2020\|EPI_ISL_543705\|2020-0 | ···················· | ·············A······ | ···················· | ···················· | NNNNNNNNNNNNNNNNNNNN | NNNNNNNNNNNNNNNNNNNN | 1 |
| H | 5118 | hCoV-19/USA/TX-HMH-4544/2020\|EPI_ISL_543707\|2020-0 | ···················· | ···················· | ···················· | ···················· | NNNNNNNNNNNNNNNNNNNN | NNNNNNNNNNNNNNNNNNNN | 1 |
| H | 5119 | hCoV-19/USA/TX-HMH-4545/2020\|EPI_ISL_543708\|2020-0 | ···················· | ···················· | ···················· | ···················· | NNNNNNNNNNNNNNNNNNNN | NNNNNNNNNNNNNNNNNNNN | 1 |
| H | 5250 | hCoV-19/Spain/CN-IBV-97006489/2020\|EPI_ISL_537703\| | ···················· | ···················· | ···················· | ···················· | NNNNNNNNNNNNNNNNNNNN | NNNNNNNNNNNNNNNNNNNN | 1 |
| H | 5251 | hCoV-19/Spain/CN-IBV-97006496/2020\|EPI_ISL_537706\| | ···················· | ···················· | ···················· | ···················· | NNNNNNNNNNNNNNNNNNNN | NNNNNNNNNNNNNNNNNNNN | 1 |
| H | 5252 | hCoV-19/Spain/CN-IBV-97006498/2020\|EPI_ISL_537708\| | ···················· | ···················· | ···················· | ···················· | NNNNNNNNNNNNNNNNNNNN | NNNNNNNNNNNNNNNNNNNN | 1 |
| H | 5253 | hCoV-19/Spain/CN-IBV-97006499/2020\|EPI_ISL_537709\| | ···················· | ···················· | ···················· | ···················· | NNNNNNNNNNNNNNNNNNNN | NNNNNNNNNNNNNNNNNNNN | 1 |
| H | 5254 | hCoV-19/Spain/CN-IBV-97006505/2020\|EPI_ISL_537713\| | ···················· | ···················· | ···················· | ···················· | NNNNNNNNNNNNNNNNNNNN | NNNNNNNNNNNNNNNNNNNN | 1 |
| H | 5323 | hCoV-19/USA/TX-HMH-4412/2020\|EPI_ISL_543600\|2020-0 | ···················· | ···················· | ···················· | ···················· | ···N··········NNN··· | NN·····NN··········· | 1 |
| H | 5325 | hCoV-19/USA/TX-HMH-4417/2020\|EPI_ISL_543603\|2020-0 | ···················· | ···················· | ···················· | ···················· | NNNNNNNNNNNNNNNNNNNN | NNNNNNNNNNNNNNNNNNNN | 1 |
| H | 5326 | hCoV-19/USA/TX-HMH-4418/2020\|EPI_ISL_543604\|2020-0 | ···················· | ···················· | ···················· | ···················· | NNNNNNNNNNNNNNNNNNNN | NNNNNNNNNNNNNNNNNNNN | 1 |
| H | 5336 | hCoV-19/USA/TX-HMH-4436/2020\|EPI_ISL_543620\|2020-0 | ···················· | ···················· | ···················· | ···················· | NNNNNNNNNNNNNNNNNNNN | NNNNNNNNNNNNNNNNNNNN | 1 |
| H | 5338 | hCoV-19/USA/TX-HMH-4442/2020\|EPI_ISL_543623\|2020-0 | ···················· | ···················· | ···················· | ···················· | NNNNNNNNNNNNNNNNNNNN | NNNNNNNNNNNNNNNNNNNN | 1 |
| H | 5339 | hCoV-19/USA/TX-HMH-4445/2020\|EPI_ISL_543625\|2020-0 | ···················· | ···················· | ···················· | ···················· | NNNNNNNNNNNNNNNNNNNN | NNNNNNNNNNNNNNNNNNNN | 1 |
| H | 5340 | hCoV-19/USA/TX-HMH-4448/2020\|EPI_ISL_543626\|2020-0 | ···················· | ···················· | ···················· | ···················· | NNNNNNNNNNNNNNNNNNNN | NNNNNNNNNNNNNNNNNNNN | 1 |
| H | 5341 | hCoV-19/USA/TX-HMH-4449/2020\|EPI_ISL_543627\|2020-0 | ···················· | ···················· | ···················· | ···················· | NNNNNNNNNNNNNNNNNNNN | NNNNNNNNNNNNNNNNNNNN | 1 |
| H | 5342 | hCoV-19/USA/TX-HMH-4450/2020\|EPI_ISL_543628\|2020-0 | ···················· | ···················· | ···················· | ···················· | NNNNNNNNNNNNNNNNNNNN | NNNNNNNNNNNNNNNNNNNN | 1 |
| H | 5343 | hCoV-19/USA/TX-HMH-4452/2020\|EPI_ISL_543629\|2020-0 | ···················· | ···················· | ···················· | ···················· | NNNNNNNNNNNNNNNNNNNN | NNNNNNNNNNNNNNNNNNNN | 1 |
| H | 5344 | hCoV-19/USA/TX-HMH-4453/2020\|EPI_ISL_543630\|2020-0 | ···················· | ···················· | ···················· | ···················· | NNNNNNNNNNNNNNNNNNNN | NNNNNNNNNNNNNNNNNNNN | 1 |
| H | 5346 | hCoV-19/USA/TX-HMH-4455/2020\|EPI_ISL_543632\|2020-0 | ···················· | ···················· | ···················· | ···················· | NNNNNNNNNNNNNNNNNNNN | NNNNNNNNNNNNNNNNNNNN | 1 |
| H | 5347 | hCoV-19/USA/TX-HMH-4456/2020\|EPI_ISL_543633\|2020-0 | ···················· | ···················· | ···················· | ···················· | NNNNNNNNNNNNNNNNNNNN | NNNNNNNNNNNNNNNNNNNN | 1 |
| H | 5348 | hCoV-19/USA/TX-HMH-4457/2020\|EPI_ISL_543634\|2020-0 | ···················· | ···················· | ···················· | ···················· | NNNNNNNNNNNNNNNNNNNN | NNNNNNNNNNNNNNNNNNNN | 1 |
| H | 5350 | hCoV-19/USA/TX-HMH-4459/2020\|EPI_ISL_543636\|2020-0 | ···················· | ···················· | ···················· | ···················· | NNNNNNNNNNNNNNNNNNNN | NNNNNNNNNNNNNNNNNNNN | 1 |
| H | 5351 | hCoV-19/USA/TX-HMH-4460/2020\|EPI_ISL_543637\|2020-0 | ···················· | ···················· | ···················· | ···················· | NNNNNNNNNNNNNNNNNNNN | NNNNNNNNNNNNNNNNNNNN | 1 |
| H | 5352 | hCoV-19/USA/TX-HMH-4461/2020\|EPI_ISL_543638\|2020-0 | ···················· | ···················· | ···················· | ···················· | NNNNNNNNNNNNNNNNNNNN | NNNNNNNNNNNNNNNNNNNN | 1 |
| H | 5353 | hCoV-19/USA/TX-HMH-4462/2020\|EPI_ISL_543639\|2020-0 | ···················· | ·············A······ | ···················· | ···················· | NNNNNNNNNNNNNNNNNNNN | NNNNNNNNNNNNNNNNNNNN | 1 |
| H | 5354 | hCoV-19/USA/TX-HMH-4463/2020\|EPI_ISL_543640\|2020-0 | ···················· | ···················· | ···················· | ···················· | NNNNNNNNNNNNNNNNNNNN | NNNNNNNNNNNNNNNNNNNN | 1 |
| H | 5356 | hCoV-19/USA/TX-HMH-4465/2020\|EPI_ISL_543642\|2020-0 | ···················· | ··············C····· | ···················· | ···················· | NNNNNNNNNNNNNNNNNNNN | NNNNNNNNNNNNNNNNNNNN | 1 |
| H | 5357 | hCoV-19/USA/TX-HMH-4466/2020\|EPI_ISL_543643\|2020-0 | ···················· | ······A············· | ···················· | ···················· | NNNNNNNNNNNNNNNNNNNN | NNNNNNNNNNNNNNNNNNNN | 1 |
| H | 5358 | hCoV-19/USA/TX-HMH-4467/2020\|EPI_ISL_543644\|2020-0 | ···················· | ···················· | ···················· | ···················· | NNNNNNNNNNNNNNNNNNNN | NNNNNNNNNNNNNNNNNNNN | 1 |
| H | 5359 | hCoV-19/USA/TX-HMH-4468/2020\|EPI_ISL_543645\|2020-0 | ···················· | ···················· | ···················· | ···················· | NNNNNNNNNNNNNNNNNNNN | NNNNNNNNNNNNNNNNNNNN | 1 |
| H | 5360 | hCoV-19/USA/TX-HMH-4470/2020\|EPI_ISL_543646\|2020-0 | ···················· | ···················· | ···················· | ···················· | NNNNNNNNNNNNNNNNNNNN | NNNNNNNNNNNNNNNNNNNN | 1 |
| H | 5361 | hCoV-19/USA/TX-HMH-4474/2020\|EPI_ISL_543650\|2020-0 | ···················· | ···················· | ···················· | ···················· | ···················· | ···N···NN··N········ | 1 |
| H | 5362 | hCoV-19/USA/TX-HMH-4476/2020\|EPI_ISL_543651\|2020-0 | ···················· | ····T··············· | ···················· | ···················· | NNNNNNNNNNNNNNNNNNNN | NNNNNNNNNNNNNNNNNNNN | 1 |
| H | 5363 | hCoV-19/USA/TX-HMH-4477/2020\|EPI_ISL_543652\|2020-0 | ···················· | ···················· | ···················· | ···················· | NNNNNNNNNNNNNNNNNNNN | NNNNNNNNNNNNNNNNNNNN | 1 |
| H | 5364 | hCoV-19/USA/TX-HMH-4478/2020\|EPI_ISL_543653\|2020-0 | ···················· | ···················· | ···················· | ···················· | NNNNNNNNNNNNNNNNNNNN | NNNNNNNNNNNNNNNNNNNN | 1 |
| H | 5365 | hCoV-19/USA/TX-HMH-4479/2020\|EPI_ISL_543654\|2020-0 | ···················· | ···················· | ···················· | ···················· | NNNNNNNNNNNNNNNNNNNN | NNNNNNNNNNNNNNNNNNNN | 1 |
| H | 5367 | hCoV-19/USA/TX-HMH-4481/2020\|EPI_ISL_543656\|2020-0 | ···················· | ···················· | ···················· | ···················· | NNNNNNNNNNNNNNNNNNNN | NNNNNNNNNNNNNNNNNNNN | 1 |
| H | 5368 | hCoV-19/USA/TX-HMH-4482/2020\|EPI_ISL_543657\|2020-0 | ···················· | ···················· | ···················· | ···················· | NNNNNNNNNNNNNNNNNNNN | NNNNNNNNNNNNNNNNNNNN | 1 |
| H | 5370 | hCoV-19/USA/TX-HMH-4485/2020\|EPI_ISL_543659\|2020-0 | ···················· | ···················· | ···················· | ···················· | NNNNNNNNNNNNNNNNNNNN | NNNNNNNNNNNNNNNNNNNN | 1 |

| | | | | | | | | | |
|---|---|---|---|---|---|---|---|---|---|
| H | 5372 | hCoV-19/USA/TX-HMH-4490/2020\|EPI_ISL_543663\|2020-0 | ···················· | ···················· | ···················· | ···················· | NNNNNNNNNNNNNNNNNNNN | NNNNNNNNNNNNNNNNNNNN | 1 |
| H | 5373 | hCoV-19/USA/TX-HMH-4493/2020\|EPI_ISL_543665\|2020-0 | ···················· | ···················· | ···················· | ···················· | N··················· | ···N········NNNN···· | 1 |
| H | 5374 | hCoV-19/USA/TX-HMH-4499/2020\|EPI_ISL_543670\|2020-0 | ···················· | N··············N···· | ···················· | ···················· | NNNNNNNNNNNNNNNNNNNN | NNNNNNNNNNNNNNNNNNNN | 1 |
| H | 5521 | hCoV-19/Spain/CL-IBV-97006240/2020\|EPI_ISL_537614\| | ···················· | ···················· | ···················· | ···················· | NNNNNNNNNNNNNNNNNNNN | NNNNNNNNNNNNNNNNNNNN | 1 |
| H | 5522 | hCoV-19/Spain/CL-IBV-97006241/2020\|EPI_ISL_537615\| | ···················· | ···················· | ···················· | ···················· | NNNNNNNNNNNNNNNNNNNN | NNNNNNNNNNNNNNNNNNNN | 1 |
| H | 5523 | hCoV-19/Spain/CL-IBV-97006247/2020\|EPI_ISL_537619\| | ···················· | ···················· | ···················· | ···················· | NNNNNNNNNNNNNNNNNNNN | NNNNNNNNNNNNNNNNNNNN | 1 |
| H | 5524 | hCoV-19/Spain/CL-IBV-97006249/2020\|EPI_ISL_537621\| | ···················· | ···················· | ···················· | ···················· | NNNNNNNNNNNNNNNNNNNN | NNNNNNNNNNNNNNNNNNNN | 1 |
| H | 5525 | hCoV-19/Spain/CL-IBV-97006252/2020\|EPI_ISL_537624\| | ···················· | ···················· | ···················· | ···················· | NNNNNNNNNNNNNNNNNNNN | ···················· | 1 |
| H | 5526 | hCoV-19/Spain/CL-IBV-97006254/2020\|EPI_ISL_537626\| | ···················· | ···················· | ···················· | ···················· | NNNNNNN···········NN | NNNNNNNNNNNNNNN····· | 1 |
| H | 5527 | hCoV-19/Spain/CL-IBV-97006259/2020\|EPI_ISL_537631\| | ···················· | ···················· | ···················· | ···················· | NNNNNNNNNNNN·NNNNNNN | ···················· | 1 |
| H | 5528 | hCoV-19/Spain/CL-IBV-97006266/2020\|EPI_ISL_537638\| | ···················· | ···················· | ···················· | ···················· | NNNNNNN············· | ···················· | 1 |
| H | 5529 | hCoV-19/Spain/CL-IBV-97006273/2020\|EPI_ISL_537644\| | ···················· | ···················· | ···················· | ···················· | NNNNNNNNNNNNNNNNNNNN | NNNNNNNNNNNNNNNNNNNN | 1 |
| H | 5530 | hCoV-19/Spain/CL-IBV-97006274/2020\|EPI_ISL_537645\| | ···················· | ···················· | ···················· | ···················· | NNNNNNNNNNNNNNNNNNNN | ···················· | 1 |
| H | 5531 | hCoV-19/Spain/CL-IBV-97006275/2020\|EPI_ISL_537646\| | ···················· | ···················· | ···················· | ···················· | NNNNNNN············· | ···················· | 1 |
| H | 5532 | hCoV-19/Spain/CL-IBV-97006280/2020\|EPI_ISL_537651\| | ···················· | ···················· | ···················· | ···················· | NNNNNNNNNNNNNNNNNNNN | NNNNNNNNNNNNNNN····· | 1 |
| H | 5533 | hCoV-19/Spain/CL-IBV-97006282/2020\|EPI_ISL_537652\| | ···················· | ···················· | ···················· | ···················· | NNNNNNNNNNNNNNNNNNNN | NNNNNNNNNNNNNNNNNNNN | 1 |
| H | 5534 | hCoV-19/Spain/CL-IBV-97006293/2020\|EPI_ISL_537663\| | ···················· | ·R·················· | ···················· | ···················· | NNNNNNNNNNNNNNNNNNNN | NNNNNNNNNNNNNNNNNNNN | 1 |
| H | 5626 | hCoV-19/USA/TX-HMH-4494/2020\|EPI_ISL_543666\|2020-0 | ···················· | ···················· | ···················· | ···················· | NNNNNNNNNNNNNNNNNNNN | NNNNNNNNNNNNNNNNNNNN | 1 |
| H | 5627 | hCoV-19/USA/TX-HMH-4495/2020\|EPI_ISL_543667\|2020-0 | ···················· | ···················· | ···················· | ···················· | NNNNNNNNNNNNNNNNNNNN | NNNNNNNNNNNNNNNNNNNN | 1 |
| H | 5631 | hCoV-19/USA/TX-HMH-4505/2020\|EPI_ISL_543675\|2020-0 | ···················· | ···················· | ···················· | ···················· | NNNNNNNNNNNNNNNNNNNN | NNNNNNNNNNNNNNNNNNNN | 1 |
| H | 5632 | hCoV-19/USA/TX-HMH-4506/2020\|EPI_ISL_543676\|2020-0 | ···················· | ···················· | ···················· | ···················· | NNNNNNNNNNNNNNNNNNNN | NNNNNNNNNNNNNNNNNNNN | 1 |
| H | 5727 | hCoV-19/Spain/CL-IBV-97006307/2020\|EPI_ISL_537676\| | ···················· | ···················· | ···················· | ···················· | NNNNNNNNNNNNNNNNNNNN | NNNNNNNNNNNNNNNNNNNN | 1 |
| H | 5841 | hCoV-19/USA/TX-HMH-4205/2020\|EPI_ISL_543436\|2020-0 | ···················· | ···················· | ···················· | ···················· | NNNNNNNNNNNNNNNNNNNN | NNNNNNNNNNNNNNNNNNNN | 1 |
| H | 5849 | hCoV-19/USA/TX-HMH-4214/2020\|EPI_ISL_543444\|2020-0 | ···················· | ···················· | ···················· | ···················· | NNNNNNNNNNNNNNNNNNNN | NNNNNNNNNNNNNNNNNNNN | 1 |
| H | 5850 | hCoV-19/USA/TX-HMH-4217/2020\|EPI_ISL_543446\|2020-0 | ···················· | ···················· | ···················· | ···················· | NNNNNNNNNNNNNNNNNNNN | NNNNNNNNNNNNNNNNNNNN | 1 |
| H | 5851 | hCoV-19/USA/TX-HMH-4218/2020\|EPI_ISL_543447\|2020-0 | ···················· | ···················· | ···················· | ···················· | NNNNNNNNNNNNNNNNNNNN | NNNNNNNNNNNNNNNNNNNN | 1 |
| H | 5852 | hCoV-19/USA/TX-HMH-4226/2020\|EPI_ISL_543450\|2020-0 | ···················· | ·A·················· | ···················· | ···················· | NNNNNNNNNNNNNNNNNNNN | NNNNNNNNNNNNNNNNNNNN | 1 |
| H | 5853 | hCoV-19/USA/TX-HMH-4239/2020\|EPI_ISL_543453\|2020-0 | ···················· | ···················· | ···················· | ···················· | NNNNNNNNNNNNNNNNNNNN | NNNNNNNNNNNNNNNNNNNN | 1 |
| H | 5854 | hCoV-19/USA/TX-HMH-4240/2020\|EPI_ISL_543454\|2020-0 | ···················· | ···················· | ···················· | ···················· | NNNNNNNNNNNNNNNNNNNN | NNNNNNNNNNNNNNNNNNNN | 1 |
| H | 5858 | hCoV-19/USA/TX-HMH-4246/2020\|EPI_ISL_543459\|2020-0 | ···················· | ···················· | ···················· | ···················· | NNNNNNNNNNNNNNNNNNNN | NNNNNNNNNNNNNNNNNNNN | 1 |
| H | 5862 | hCoV-19/USA/TX-HMH-4253/2020\|EPI_ISL_543465\|2020-0 | ···················· | ···················· | ···················· | ···················· | NNNNNNNNNNNNNNNNNNNN | NNNNNNNNNNNNNNNNNNNN | 1 |
| H | 5863 | hCoV-19/USA/TX-HMH-4254/2020\|EPI_ISL_543466\|2020-0 | ···················· | ···················· | ···················· | ···················· | NNNNNNNNNNNNNNNNNNNN | NNNNNNNNNNNNNNNNNNNN | 1 |
| H | 5864 | hCoV-19/USA/TX-HMH-4255/2020\|EPI_ISL_543467\|2020-0 | ···················· | ···················· | ···················· | ···················· | ········N···N·NN···· | NNNNNNNNN···NNN····N | 1 |
| H | 5871 | hCoV-19/USA/TX-HMH-4264/2020\|EPI_ISL_543476\|2020-0 | ···················· | NNNNNNNNNNNNNNNNNNNN | ···················· | ···················· | ···················· | ···················· | 1 |
| H | 5875 | hCoV-19/USA/TX-HMH-4269/2020\|EPI_ISL_543481\|2020-0 | ···················· | ·A·················· | ···················· | ···················· | NNNNNNNNNNNNNNNNNNNN | NNNNNNNNNNNNNNNNNNNN | 1 |
| H | 5876 | hCoV-19/USA/TX-HMH-4271/2020\|EPI_ISL_543483\|2020-0 | ···················· | ···················· | ···················· | ···················· | NNNNNNNNNNNNNNNNNNNN | NNNNNNNNNNNNNNNNNNNN | 1 |
| H | 5877 | hCoV-19/USA/TX-HMH-4273/2020\|EPI_ISL_543485\|2020-0 | ···················· | ···················· | ···················· | ···················· | NNNNNNNNNNNNNNNNNNNN | NNNNNNNNNNNNNNNNNNNN | 1 |
| H | 5880 | hCoV-19/USA/TX-HMH-4276/2020\|EPI_ISL_543488\|2020-0 | ···················· | N·················N· | ···················· | ···················· | NNNNNNNNNNNNNNNNNNNN | NNNNNNNNNNNNNNNNNNNN | 1 |
| H | 5881 | hCoV-19/USA/TX-HMH-4277/2020\|EPI_ISL_543489\|2020-0 | ···················· | ···················· | ···················· | ···················· | NNNNNNNNNNNNNNNNNNNN | NNNNNNNNNNNNNNNNNNNN | 1 |
| H | 6075 | hCoV-19/Spain/CN-IBV-97006482/2020\|EPI_ISL_537697\| | ···················· | ···················· | ···················· | ···················· | NNNNNNNNNNNNNNNNNNNN | NNNNNNNNNNNNNNNNNNNN | 1 |
| H | 6103 | hCoV-19/USA/TX-HMH-4293/2020\|EPI_ISL_543503\|2020-0 | ···················· | ···················· | ···················· | ···················· | ·····NNN······N····N | N······NN··········· | 1 |
| H | 6111 | hCoV-19/USA/TX-HMH-4304/2020\|EPI_ISL_543514\|2020-0 | ···················· | ···················· | ···················· | ···················· | NNNNNNNNNNNNNNNNNNNN | NNNNNNNNNNNNNNNNNNNN | 1 |
| H | 6115 | hCoV-19/USA/TX-HMH-4311/2020\|EPI_ISL_543520\|2020-0 | ···················· | N············A······ | ···················· | ···················· | NNNNNNNNNNNNNNNNNNNN | NNNNNNNNNNNNNNNNNNNN | 1 |
| H | 6117 | hCoV-19/USA/TX-HMH-4314/2020\|EPI_ISL_543523\|2020-0 | ···················· | ···················· | ···················· | ···················· | NNNNNNNNNNNNNNNNNNNN | NNNNNNNNNNNNNNNNNNNN | 1 |
| H | 6121 | hCoV-19/USA/TX-HMH-4319/2020\|EPI_ISL_543528\|2020-0 | ···················· | ···················· | ···················· | ···················· | NNNNNNNNNNNNNNNNNNNN | NNNNNNNNNNNNNNNNNNNN | 1 |
| H | 6124 | hCoV-19/USA/TX-HMH-4323/2020\|EPI_ISL_543532\|2020-0 | ···················· | ·············A······ | ···················· | ···················· | NNNNNNNNNNNNNNNNNNNN | NNNNNNNNNNNNNNNNNNNN | 1 |
| H | 6127 | hCoV-19/USA/TX-HMH-4326/2020\|EPI_ISL_543535\|2020-0 | ···················· | ···················· | ···················· | ···················· | NNNNNNNNNNNNNNNNNNNN | NNNNNNNNNNNNNNNNNNNN | 1 |
| H | 6128 | hCoV-19/USA/TX-HMH-4327/2020\|EPI_ISL_543536\|2020-0 | ···················· | ···················· | ···················· | ···················· | ········N··········· | NN·N···NN·····N····· | 1 |
| H | 6133 | hCoV-19/USA/TX-HMH-4339/2020\|EPI_ISL_543543\|2020-0 | ···················· | ···················· | ···················· | ···················· | NNNNNNNNNNNNNNNNNNNN | NNNNNNNNNNNNNNNNNNNN | 1 |
| H | 6143 | hCoV-19/USA/TX-HMH-4353/2020\|EPI_ISL_543554\|2020-0 | ···················· | ···················· | ···················· | ···················· | NNNNNNNNNNNNNNNNNNNN | NNNNNNNNNNNNNNNNNNNN | 1 |
| H | 6384 | hCoV-19/USA/TX-HMH-4037/2020\|EPI_ISL_543316\|2020-0 | ···················· | ···················· | ···················· | ···················· | NNNNNNNNNNNNNNNNNNNN | NNNNNNNNNNNNNNNNNNNN | 1 |
| H | 6394 | hCoV-19/USA/TX-HMH-4051/2020\|EPI_ISL_543327\|2020-0 | ···················· | ···················· | ···················· | ···················· | NNNNNNNNNNNNNNNNNNNN | NNNNNNNNNNNNNNNNNNNN | 1 |
| H | 6599 | hCoV-19/USA/TX-HMH-4070/2020\|EPI_ISL_543342\|2020-0 | ···················· | ···················· | ···················· | ···················· | NNNNNNNNNNNNNNNNNNNN | NNNNNNNNNNNNNNNNNNNN | 1 |
| H | 6601 | hCoV-19/USA/TX-HMH-4074/2020\|EPI_ISL_543345\|2020-0 | ···················· | ···················· | ···················· | ···················· | ··············N····· | NN·N···NN····NN····· | 1 |
| H | 6604 | hCoV-19/USA/TX-HMH-4080/2020\|EPI_ISL_543351\|2020-0 | ···················· | ···················· | ···················· | ···················· | NNNNNNNNNNNNNNNNNNNN | NNNNNNNNNNNNNNNNNNNN | 1 |
| H | 6605 | hCoV-19/USA/TX-HMH-4082/2020\|EPI_ISL_543353\|2020-0 | ···················· | ···················· | ···················· | ···················· | NNNNNNNNNNNNNNNNNNNN | NNNNNNNNNNNNNNNNNNNN | 1 |
| H | 6607 | hCoV-19/USA/TX-HMH-4086/2020\|EPI_ISL_543356\|2020-0 | ···················· | ···················· | ···················· | ···················· | NNNNNNNNNNNNNNNNNNNN | NNNNNNNNNNNNNNNNNNNN | 1 |
| H | 6608 | hCoV-19/USA/TX-HMH-4087/2020\|EPI_ISL_543357\|2020-0 | ···················· | ···················· | ···················· | ···················· | ··N·····NN····NNNN·· | NN·NN··NN·NN········ | 1 |
| H | 6613 | hCoV-19/USA/TX-HMH-4093/2020\|EPI_ISL_543363\|2020-0 | ···················· | ·············A······ | ···················· | ···················· | ·N·N··········NN·N·· | NNNN···NN··········· | 1 |
| H | 6622 | hCoV-19/USA/TX-HMH-4109/2020\|EPI_ISL_543377\|2020-0 | ···················· | ···················· | ···················· | ···················· | NNNNNNNNNNNNNNNNNNNN | NNNNNNNNNNNNNNNNNNNN | 1 |
| H | 6640 | hCoV-19/USA/TX-HMH-4141/2020\|EPI_ISL_543404\|2020-0 | ···················· | ·············A······ | ···················· | ···················· | NNNNNNNNNNNNNNNNNNNN | NNNNNNNNNNNNNNNNNNNN | 1 |
| H | 6644 | hCoV-19/USA/TX-HMH-4169/2020\|EPI_ISL_543408\|2020-0 | ···················· | ···················· | ···················· | ···················· | NNNNNNNNNNNNNNNNNNNN | NNNNNNNNNNNNNNNNNNNN | 1 |
| H | 6646 | hCoV-19/USA/TX-HMH-4173/2020\|EPI_ISL_543410\|2020-0 | ···················· | ···················· | ···················· | ···················· | NN·N·········NN··N·· | NN·NN··NNN····N····· | 1 |
| H | 6647 | hCoV-19/USA/TX-HMH-4174/2020\|EPI_ISL_543411\|2020-0 | ···················· | ·A·················· | ···················· | ···················· | ··············N····· | NN······N·NN········ | 1 |
| H | 6648 | hCoV-19/USA/TX-HMH-4175/2020\|EPI_ISL_543412\|2020-0 | ···················· | ···················· | ···················· | ···················· | NNNNNNNNNNNNNNNNNNNN | NNNNNNNNNNNNNNNNNNNN | 1 |
| H | 6649 | hCoV-19/USA/TX-HMH-4176/2020\|EPI_ISL_543413\|2020-0 | ···················· | ···················· | ···················· | ···················· | ········NN····N····· | NN·N···NNN·····N···· | 1 |
| H | 6655 | hCoV-19/USA/TX-HMH-4188/2020\|EPI_ISL_543422\|2020-0 | ···················· | ·············A······ | ···················· | ···················· | NNNNNNNNNNNNNNNNNNNN | NNNNNNNNNNNNNNNNNNNN | 1 |
| H | 6656 | hCoV-19/USA/TX-HMH-4189/2020\|EPI_ISL_543423\|2020-0 | ···················· | ···················· | ···················· | ···················· | NNNNNNNNNNNNNNNNNNNN | NNNNNNNNNNNNNNNNNNNN | 1 |
| H | 6906 | hCoV-19/USA/TX-HMH-4191/2020\|EPI_ISL_543425\|2020-0 | ···················· | ···················· | ···················· | ···················· | NNNNNNNNNNNNNNNNNNNN | NNNNNNNNNNNNNNNNNNNN | 1 |
| H | 7243 | hCoV-19/Spain/VC-IBV-98003447/2020\|EPI_ISL_539256\| | ···················· | ···················· | ···················· | ···················· | ··NNNNN············· | ···················· | 1 |
| H | 7244 | hCoV-19/Spain/VC-IBV-98003450/2020\|EPI_ISL_539257\| | ···················· | ···················· | ···················· | ···················· | ·NNNNNNNNNNNNNNNNNNN | NNNNNNNNNNNNNNN··NNN | 1 |
| H | 7251 | hCoV-19/Spain/VC-IBV-98003496/2020\|EPI_ISL_539286\| | ···················· | ···················· | ···················· | ···················· | NNNNNNNNNNNNNNNNNNNN | NNNNNNNNNNNNNNNNNNNN | 1 |
| H | 7252 | hCoV-19/Spain/VC-IBV-98003497/2020\|EPI_ISL_539287\| | ···················· | ···················· | ···················· | ···················· | NNNNNNNNNNNNNNNNNNNN | NNNNNNNNNNNNNNNNNNNN | 1 |
| H | 7323 | hCoV-19/USA/IL-UW-1518/2020\|EPI_ISL_541547\|2020-09 | ···················· | ···················· | ···················· | ···················· | NNNNNNNNNNNNNNNNNNNN | NNNNNNNNNNNNNNNNNNNN | 1 |
| H | 7328 | hCoV-19/USA/WI-UW-1419/2020\|EPI_ISL_541552\|2020-09 | ···················· | ···················· | ···················· | ···················· | NNNNNNNNNNNNNNNNNNNN | NNNNNNNNNNNNNNNNNNNN | 1 |
| H | 7329 | hCoV-19/USA/WI-UW-1420/2020\|EPI_ISL_541553\|2020-09 | ···················· | ···················· | ···················· | ···················· | NNNNNNNNNNNNNNNNNNNN | NNNNNNNNNNNNNNNNNNNN | 1 |
| H | 7332 | hCoV-19/USA/WI-UW-1423/2020\|EPI_ISL_541556\|2020-09 | ···················· | ···················· | ···················· | ···················· | NNNNNNNNNNNNNNNNNNNN | NNNNNNNNNNNNNNNNNNNN | 1 |
| H | 7333 | hCoV-19/USA/WI-UW-1424/2020\|EPI_ISL_541557\|2020-09 | ···················· | ···················· | ···················· | ···················· | NNNNNNNNNNNNNNNNNNNN | NNNNNNNNNNNNNNNNNNNN | 1 |
| H | 7334 | hCoV-19/USA/WI-UW-1425/2020\|EPI_ISL_541558\|2020-09 | ···················· | ···················· | ···················· | ···················· | NNNNNNNNNNNNNNNNNNNN | NNNNNNNNNNNNNNNNNNNN | 1 |
| H | 7337 | hCoV-19/USA/WI-UW-1428/2020\|EPI_ISL_541561\|2020-09 | ···················· | ···················· | ···················· | ···················· | NNNNNNNNNNNNNNNNNNNN | NNNNNNNNNNNNNNNNNNNN | 1 |

| | | | | | | | | | |
|---|---|---|---|---|---|---|---|---|---|
| H | 7339 | hCoV-19/USA/WI-UW-1430/2020\|EPI_ISL_541563\|2020-09 | ···················· | ···················· | ···················· | ···················· | NNNNNNNNNNNNNNNNNNNN | NNNNNNNNNNNNNNNNNNNN | 1 |
| H | 7341 | hCoV-19/USA/WI-UW-1432/2020\|EPI_ISL_541565\|2020-09 | ···················· | ···················· | ···················· | ···················· | NNNNNNNNNNNNNNNNNNNN | NNNNNNNNNNNNNNNNNNNN | 1 |
| H | 7346 | hCoV-19/USA/WI-UW-1438/2020\|EPI_ISL_541571\|2020-09 | ···················· | ···················· | ···················· | ···················· | NNNNNNNNNNNNNNNNNNNN | NNNNNNNNNNNNNNNNNNNN | 1 |
| H | 7347 | hCoV-19/USA/WI-UW-1441/2020\|EPI_ISL_541573\|2020-09 | ···················· | ···················· | ···················· | ···················· | NNNNNNNNNNNNNNNNNNNN | NNNNNNNNNNNNNNNNNNNN | 1 |
| H | 7351 | hCoV-19/USA/WI-UW-1445/2020\|EPI_ISL_541577\|2020-09 | ···················· | ···················· | ···················· | ···················· | NNNNNNNNNNNNNNNNNNNN | NNNNNNNNNNNNNNNNNNNN | 1 |
| H | 7352 | hCoV-19/USA/WI-UW-1446/2020\|EPI_ISL_541578\|2020-09 | ···················· | ···················· | ···················· | ···················· | NNNNNNNNNNNNNNNNNNNN | NNNNNNNNNNNNNNNNNNNN | 1 |
| H | 7358 | hCoV-19/USA/WI-UW-1452/2020\|EPI_ISL_541584\|2020-09 | ···················· | ···················· | ···················· | ···················· | NNNNNNNNNNNNNNNNNNNN | NNNNNNNNNNNNNNNNNNNN | 1 |
| H | 7363 | hCoV-19/USA/WI-UW-1457/2020\|EPI_ISL_541589\|2020-09 | ···················· | ···················· | ···················· | ···················· | NNNNNNNNNNNNNNNNNNNN | NNNNNNNNNNNNNNNNNNNN | 1 |
| H | 7365 | hCoV-19/USA/WI-UW-1459/2020\|EPI_ISL_541591\|2020-09 | ···················· | ···················· | ···················· | ···················· | NNNNNNNNNNNNNNNNNNNN | NNNNNNNNNNNNNNNNNNNN | 1 |
| H | 7370 | hCoV-19/USA/WI-UW-1464/2020\|EPI_ISL_541596\|2020-09 | ···················· | ···················· | ···················· | ···················· | NNNNNNNNNNNNNNNNNNNN | NNNNNNNNNNNNNNNNNNNN | 1 |
| H | 7371 | hCoV-19/USA/WI-UW-1465/2020\|EPI_ISL_541597\|2020-09 | ···················· | ···················· | ···················· | ···················· | NNNNNNNNNNNNNNNNNNNN | NNNNNNNNNNNNNNNNNNNN | 1 |
| H | 7378 | hCoV-19/USA/TX-HMH-1737/2020\|EPI_ISL_542804\|2020-0 | ···················· | ···················· | ···················· | ···················· | NNNNNNNNNNNNNNNNNNNN | NNNNNNNNNNNNNNNNNNNN | 1 |
| H | 7391 | hCoV-19/USA/TX-HMH-1759/2020\|EPI_ISL_542819\|2020-0 | ···················· | ···················· | ···················· | ···················· | NNNNNNNNNNNNNNNNNNNN | NNNNNNNNNNNNNNNNNNNN | 1 |
| H | 7663 | hCoV-19/USA/TX-HMH-1793/2020\|EPI_ISL_542842\|2020-0 | ···················· | ·············C······ | ···················· | ···················· | NNNNNNNNNNNNNNNNNNNN | NNNNNNNNNNNNNNNNNNNN | 1 |
| H | 7670 | hCoV-19/USA/TX-HMH-1800/2020\|EPI_ISL_542849\|2020-0 | ···················· | ·············C······ | ···················· | ···················· | ··NN··········N····· | NN····NN···N········ | 1 |
| H | 7671 | hCoV-19/USA/TX-HMH-1802/2020\|EPI_ISL_542850\|2020-0 | ···················· | ·····A·············· | ···················· | ···················· | NNNNNNNNNNNNNNNNNNNN | NNNNNNNNNNNNNNNNNNNN | 1 |
| H | 7922 | hCoV-19/USA/TX-HMH-1496/2020\|EPI_ISL_542754\|2020-0 | ···················· | ···················· | ···················· | ···················· | NNNNNNNNNNNNNNNNNNNN | ···················· | 1 |
| H | 7928 | hCoV-19/USA/TX-HMH-1506/2020\|EPI_ISL_542761\|2020-0 | ···················· | ···················· | ···················· | ···················· | NNNNNNNNNNNNNNNNNNNN | ··NNNNNNN···NNNNNNNN | 1 |
| H | 8152 | hCoV-19/USA/TX-HMH-1432/2020\|EPI_ISL_542720\|2020-0 | ···················· | ···················· | ···················· | ···················· | NNNNNNNNNNNNNNNNNNNN | ···················· | 1 |
| H | 8165 | hCoV-19/USA/TX-HMH-1459/2020\|EPI_ISL_542734\|2020-0 | ···················· | ···················· | ···················· | ···················· | NNNNNNNNNNNNNNNNNNNN | ···················· | 1 |
| H | 8292 | hCoV-19/South Africa/KRISP-K002614/2020\|EPI_ISL_53 | ···················· | ···················· | ···················· | ···················· | NNNNNNNNNNNN········ | ···················· | 1 |
| H | 8315 | hCoV-19/USA/WI-UW-1339/2020\|EPI_ISL_536710\|2020-09 | ···················· | ···················· | ···················· | ···················· | NNNNNNNNNNNNNNNNNNNN | NNNNNNNNNNNNNNNNNNNN | 1 |
| H | 8318 | hCoV-19/USA/WI-UW-1342/2020\|EPI_ISL_536713\|2020-09 | ···················· | ···················· | ···················· | ···················· | NNNNNNNNNNNNNNNNNNNN | NNNNNNNNNNNNNNNNNNNN | 1 |
| H | 8354 | hCoV-19/USA/WI-UW-1383/2020\|EPI_ISL_536754\|2020-09 | ···················· | ···················· | ···················· | ···················· | NNNNNNNNNNNNNNNNNNNN | NNNNNNNNNNNNNNNNNNNN | 1 |
| H | 8390 | hCoV-19/USA/FL-BPHL-1324/2020\|EPI_ISL_541262\|2020- | ···················· | ···················· | ···················· | ···················· | NNNNNNNNNNNNNNNNNNNN | ···················· | 1 |
| H | 8391 | hCoV-19/USA/FL-BPHL-1325/2020\|EPI_ISL_541263\|2020- | ···················· | ···················· | ···················· | ···················· | NNNNNNNNNNNNNNNNNNNN | NNNNNNNNNNNNNNNNNNNN | 1 |
| H | 8394 | hCoV-19/USA/FL-BPHL-1328/2020\|EPI_ISL_541266\|2020- | ···················· | ···················· | ···················· | ···················· | NNNNNNNNNNNNNNNNNNNN | NNNNNNNNNNNNNNNNNNNN | 1 |
| H | 8396 | hCoV-19/USA/FL-BPHL-1330/2020\|EPI_ISL_541268\|2020- | ···················· | ···················· | ···················· | ···················· | NNNNNNNNNNNNNNNNNNNN | ···················· | 1 |
| H | 8398 | hCoV-19/USA/FL-BPHL-1333/2020\|EPI_ISL_541271\|2020- | ···················· | ···················· | ···················· | ···················· | NNNNNNNNNNNNNNNNNN·· | NNNNNNNNNNNNNNN····· | 1 |
| H | 8400 | hCoV-19/USA/FL-BPHL-1339/2020\|EPI_ISL_541277\|2020- | ···················· | ···················· | ···················· | ···················· | N··········NNNNNNNNN | NNNNNNNNNNNNNNN····· | 1 |
| H | 8411 | hCoV-19/USA/FL-BPHL-1357/2020\|EPI_ISL_541294\|2020- | ···················· | ···················· | ···················· | ···················· | NNNNNNNNNNNNNNNNNNNN | ··········NNN······· | 1 |
| H | 8412 | hCoV-19/USA/FL-BPHL-1358/2020\|EPI_ISL_541295\|2020- | ···················· | ···················· | ···················· | ···················· | NNNNNNNNNNNNNNNNNNNN | NNNNNNNNNNNNNNNNNNNN | 1 |
| H | 8413 | hCoV-19/USA/FL-BPHL-1360/2020\|EPI_ISL_541297\|2020- | ···················· | ···················· | ···················· | ···················· | NNNNNNNNNNNNNNNNNNNN | NNNNNNNNNNNNNNN····· | 1 |
| H | 8414 | hCoV-19/USA/FL-BPHL-1362/2020\|EPI_ISL_541299\|2020- | ···················· | ···················· | ···················· | ···················· | NNNNNNNNNNNNNNNNNNNN | NNNNNNNNNNNNNNNNNNNN | 1 |
| H | 8415 | hCoV-19/USA/FL-BPHL-1364/2020\|EPI_ISL_541301\|2020- | ···················· | ···················· | ···················· | ···················· | NNNNNNNNNNNNNNNNNNNN | ···················· | 1 |
| H | 8416 | hCoV-19/USA/FL-BPHL-1365/2020\|EPI_ISL_541302\|2020- | ···················· | ···················· | ···················· | ···················· | NNNNNNNNNNNNNNNNNNNN | NNNNNNNNNNNNNNNNNNNN | 1 |
| H | 8444 | hCoV-19/USA/TX-HMH-1302/2020\|EPI_ISL_542638\|2020-0 | ···················· | ···················· | ···················· | ···················· | NNNNNNNNNNNNNNNNNNNN | NNNNNNNNNNNNNNNNNNNN | 1 |
| H | 8447 | hCoV-19/USA/TX-HMH-1306/2020\|EPI_ISL_542641\|2020-0 | ···················· | ···················· | ···················· | ···················· | ···········N··N····· | NNNN···NN··········· | 1 |
| H | 8670 | hCoV-19/USA/TX-HMH-1341/2020\|EPI_ISL_542659\|2020-0 | ···················· | ···················· | ···················· | ···················· | NNNNNNNNNNNNNNNNNNNN | NNNNNNNNNNNNNNNNNNNN | 1 |
| H | 8673 | hCoV-19/USA/TX-HMH-1350/2020\|EPI_ISL_542664\|2020-0 | ···················· | ···················· | ···················· | ···················· | NNNNNNNNNNNNNNNNNNNN | NNNNNNNNNNNNNNNNNNNN | 1 |
| H | 8686 | hCoV-19/USA/TX-HMH-1372/2020\|EPI_ISL_542679\|2020-0 | ···················· | ···················· | ···················· | ···················· | NNNNNNNNNNNNNNNNNNNN | NNNNNNNNNNNNNNNNNNNN | 1 |
| H | 8889 | hCoV-19/USA/WI-UW-1307/2020\|EPI_ISL_536679\|2020-08 | ···················· | ···················· | ···················· | ···················· | NNNNNNNNNNNNNNNNNNNN | NNNNNNNNNNNNNNNNNNNN | 1 |
| H | 8890 | hCoV-19/USA/WI-UW-1308/2020\|EPI_ISL_536680\|2020-09 | ···················· | ···················· | ···················· | ···················· | NNNNNNNNNNNNNNNNNNNN | NNNNNNNNNNNNNNNNNNNN | 1 |
| H | 8891 | hCoV-19/USA/WI-UW-1309/2020\|EPI_ISL_536681\|2020-08 | ···················· | ···················· | ···················· | ···················· | NNNNNNNNNNNNNNNNNNNN | NNNNNNNNNNNNNNNNNNNN | 1 |
| H | 8892 | hCoV-19/USA/WI-UW-1310/2020\|EPI_ISL_536682\|2020-08 | ···················· | ···················· | ···················· | ···················· | NNNNNNNNNNNNNNNNNNNN | NNNNNNNNNNNNNNNNNNNN | 1 |
| H | 8893 | hCoV-19/USA/NV-UW-1311/2020\|EPI_ISL_536683\|2020-08 | ···················· | ···················· | ···················· | ···················· | NNNNNNNNNNNNNNNNNNNN | NNNNNNNNNNNNNNNNNNNN | 1 |
| H | 8894 | hCoV-19/USA/WI-UW-1313/2020\|EPI_ISL_536685\|2020-08 | ···················· | ···················· | ···················· | ···················· | NNNNNNNNNNNNNNNNNNNN | NNNNNNNNNNNNNNNNNNNN | 1 |
| H | 8896 | hCoV-19/USA/WI-UW-1315/2020\|EPI_ISL_536687\|2020-08 | ···················· | ···················· | ···················· | ···················· | NNNNNNNNNNNNNNNNNNNN | NNNNNNNNNNNNNNNNNNNN | 1 |
| H | 8897 | hCoV-19/USA/WI-UW-1316/2020\|EPI_ISL_536688\|2020-08 | ···················· | ···················· | ···················· | ···················· | NNNNNNNNNNNNNNNNNNNN | NNNNNNNNNNNNNNNNNNNN | 1 |
| H | 8898 | hCoV-19/USA/WI-UW-1317/2020\|EPI_ISL_536689\|2020-08 | ···················· | ···················· | ···················· | ···················· | NNNNNNNNNNNNNNNNNNNN | NNNNNNNNNNNNNNNNNNNN | 1 |
| H | 8899 | hCoV-19/USA/WI-UW-1323/2020\|EPI_ISL_536694\|2020-08 | ···················· | ···················· | ···················· | ···················· | NNNNNNNNNNNNNNNNNNNN | NNNNNNNNNNNNNNNNNNNN | 1 |
| H | 8905 | hCoV-19/USA/FL-BPHL-1254/2020\|EPI_ISL_541150\|2020- | ···················· | ···················· | ···················· | ···················· | NNNNNNNNNNNNNNNNNNNN | NNNNNNNNNNNNNNNNNNNN | 1 |
| H | 8906 | hCoV-19/USA/FL-BPHL-1236/2020\|EPI_ISL_541154\|2020- | ···················· | ···················· | ···················· | ···················· | NNNNNNNNNNNNNNNNNNNN | NNNNNNNNNNNNNNNNNNNN | 1 |
| H | 8911 | hCoV-19/USA/FL-BPHL-1266/2020\|EPI_ISL_541173\|2020- | ···················· | ···················· | ···················· | ···················· | NNNNNNNNNNNNNNNNNNNN | ···················· | 1 |
| H | 8912 | hCoV-19/USA/FL-BPHL-1267/2020\|EPI_ISL_541174\|2020- | ···················· | ···················· | ···················· | ···················· | ··NNNNNNNNNNNNNNNNNN | ···················· | 1 |
| H | 8917 | hCoV-19/USA/FL-BPHL-1207/2020\|EPI_ISL_541180\|2020- | ···················· | ···················· | ···················· | ···················· | NNNNNNNNNNNNNNNNNNNN | NNNNNNNNNNNNNNNNNNNN | 1 |
| H | 8927 | hCoV-19/USA/FL-BPHL-1220/2020\|EPI_ISL_541193\|2020- | ···················· | ···················· | ···················· | ···················· | NNNNNNNNNNNNNNNNNNNN | NNNNNNNNNNNNNNNNNNNN | 1 |
| H | 8930 | hCoV-19/USA/FL-BPHL-1225/2020\|EPI_ISL_541198\|2020- | ···················· | ···················· | ···················· | ···················· | NNNNNNNNNNNNNNNNNNNN | ···················· | 1 |
| H | 8957 | hCoV-19/USA/TX-HMH-1257/2020\|EPI_ISL_542616\|2020-0 | ···················· | ···················· | ···················· | ···················· | NNNNNNNNNNNNNNNNNNNN | NNNNNNNNNNNNNNNNNNNN | 1 |
| H | 9102 | hCoV-19/USA/WI-UW-1246/2020\|EPI_ISL_536619\|2020-08 | ···················· | ···················· | ···················· | ···················· | NNNNNNNNNNNNNNNNNNNN | NNNNNNNNNNNNNNNNNNNN | 1 |
| H | 9117 | hCoV-19/USA/FL-BPHL-1287/2020\|EPI_ISL_541219\|2020- | ···················· | ···················· | ···················· | ···················· | NNNNNNNNNNNNNNN····· | ···················· | 1 |
| H | 9118 | hCoV-19/USA/FL-BPHL-1288/2020\|EPI_ISL_541220\|2020- | ···················· | ···················· | ···················· | ···················· | NNNNNNNNNNNNNNNNNNNN | NNNNNNNNNNNNNNNNNNNN | 1 |
| H | 9120 | hCoV-19/USA/FL-BPHL-1290/2020\|EPI_ISL_541222\|2020- | ···················· | ···················· | ···················· | ···················· | NNNNNNNNNNNNNNNNNNNN | ···················· | 1 |
| H | 9123 | hCoV-19/USA/FL-BPHL-1299/2020\|EPI_ISL_541231\|2020- | ···················· | ···················· | ···················· | ···················· | NNNNNNNNNNNNNNNNNNNN | ···················· | 1 |
| H | 9124 | hCoV-19/USA/FL-BPHL-1300/2020\|EPI_ISL_541232\|2020- | ···················· | ···················· | ···················· | ···················· | NNNNNNNNNNNNNNNNNNNN | NNNNNNNNNNNNNNNNNNNN | 1 |
| H | 9127 | hCoV-19/USA/FL-BPHL-1304/2020\|EPI_ISL_541236\|2020- | ···················· | ···················· | ···················· | ···················· | NNNNNNNNNNNNNNNNNNNN | ···················· | 1 |
| H | 9160 | hCoV-19/USA/TX-HMH-1080/2020\|EPI_ISL_542523\|2020-0 | ···················· | ···················· | ···················· | ···················· | NNNNNNNNNNNNNNNNNNNN | NNNNNNNNNNNNNNNNNNNN | 1 |
| H | 9165 | hCoV-19/USA/TX-HMH-1096/2020\|EPI_ISL_542529\|2020-0 | ···················· | ···················· | ···················· | ···················· | ··············NN···· | NN····NN············ | 1 |
| H | 9167 | hCoV-19/USA/TX-HMH-1102/2020\|EPI_ISL_542531\|2020-0 | ···················· | ···················· | ···················· | ···················· | NNNNNNNNNNNNNNNNNNNN | ···················· | 1 |
| H | 9174 | hCoV-19/USA/TX-HMH-1112/2020\|EPI_ISL_542538\|2020-0 | ···················· | ···················· | ···················· | ···················· | NNNNNNNNNNNNNNNNNNNN | ···················· | 1 |
| H | 9176 | hCoV-19/USA/TX-HMH-1114/2020\|EPI_ISL_542540\|2020-0 | ···················· | ···················· | ···················· | ···················· | NNNNNNNNNNNNNNNNNNNN | ···················· | 1 |
| H | 9177 | hCoV-19/USA/TX-HMH-1119/2020\|EPI_ISL_542541\|2020-0 | ···················· | ···················· | ···················· | ···················· | NNNNNNNNNNNNNNNNNNNN | ···················· | 1 |
| H | 9181 | hCoV-19/USA/TX-HMH-1136/2020\|EPI_ISL_542545\|2020-0 | ···················· | ···················· | ···················· | ···················· | NNNNNNNNNNNNNNNNNNNN | ···················· | 1 |
| H | 9184 | hCoV-19/USA/TX-HMH-1140/2020\|EPI_ISL_542548\|2020-0 | ···················· | ···················· | ···················· | ···················· | NNNNNNNNNNNNNNNNNNNN | ···················· | 1 |
| H | 9186 | hCoV-19/USA/TX-HMH-1143/2020\|EPI_ISL_542550\|2020-0 | ···················· | ···················· | ···················· | ···················· | NNNNNNNNNNNNNNNNNNNN | ···················· | 1 |
| H | 9188 | hCoV-19/USA/TX-HMH-1147/2020\|EPI_ISL_542552\|2020-0 | ···················· | ···················· | ···················· | ···················· | NNNNNNNNNNNNNNNNNNNN | ···················· | 1 |
| H | 9192 | hCoV-19/USA/TX-HMH-1159/2020\|EPI_ISL_542558\|2020-0 | ···················· | ···················· | ···················· | ···················· | NNNNNNNNNNNNNNNNNNNN | NNNNNNNNNNNNNNNNNNNN | 1 |
| H | 9193 | hCoV-19/USA/TX-HMH-1161/2020\|EPI_ISL_542560\|2020-0 | ···················· | ···················· | ···················· | ···················· | NNNNNNNNNNNNNNNNNNNN | ···················· | 1 |
| H | 9197 | hCoV-19/USA/TX-HMH-1178/2020\|EPI_ISL_542567\|2020-0 | ···················· | ···················· | ···················· | ···················· | ···N·NNN·N····NN···· | NN·····N·····NN··NNN | 1 |
| I | 1 | hCoV-19/USA/TX-HMH-3318/2020\|EPI_ISL_544128\|2020-0 | ···················· | ···················· | ···················· | ···················· | NNNNNNNNNNNNNNNNNNNN | NNNNNNNNNNNNNNNNNNNN | 1 |

| | | | | | | | | | |
|---|---|---|---|---|---|---|---|---|---|
| I | 11 | hCoV-19/USA/TX-HMH-3358/2020\|EPI_ISL_544143\|2020-0 | ···················· | ···················· | ···················· | ···················· | NNNNNNNNNNNNNNNNNNNN | NNNNNNNNNNNNNNNNNNNN | 1 |
| I | 24 | hCoV-19/USA/TX-HMH-3399/2020\|EPI_ISL_544159\|2020-0 | ···················· | ···················· | ···················· | ···················· | NNNNNNNNNNNNNNNNNNNN | NNNNNNNNNNNNNNNNNNNN | 1 |
| I | 28 | hCoV-19/USA/TX-HMH-3403/2020\|EPI_ISL_544163\|2020-0 | ···················· | ···················· | ···················· | ···················· | NNNNNNNNNNNNNNNNNNNN | NNNNNNNNNNNNNNNNNNNN | 1 |
| I | 33 | hCoV-19/USA/TX-HMH-3409/2020\|EPI_ISL_544169\|2020-0 | ···················· | ···················· | ···················· | ···················· | ············N·N····· | ··NN···N·····N·N···· | 1 |
| I | 34 | hCoV-19/USA/TX-HMH-3410/2020\|EPI_ISL_544170\|2020-0 | ···················· | ···················· | ···················· | ···················· | NNNNNNNNNNNNNNNNNNNN | NNNNNNNNNNNNNNNNNNNN | 1 |
| I | 35 | hCoV-19/USA/TX-HMH-3411/2020\|EPI_ISL_544171\|2020-0 | ···················· | ···················· | ···················· | ···················· | NNNNNNNNNNNNNNNNNNNN | NNNNNNNNNNNNNNNNNNNN | 1 |
| I | 37 | hCoV-19/USA/TX-HMH-3415/2020\|EPI_ISL_544174\|2020-0 | ···················· | ···················· | ···················· | ···················· | NNNNNNNNNNNNNNNNNNNN | NNNNNNNNNNNNNNNNNNNN | 1 |
| I | 38 | hCoV-19/USA/TX-HMH-3416/2020\|EPI_ISL_544175\|2020-0 | ···················· | ···················· | ···················· | ···················· | NNNNNNNNNNNNNNNNNNNN | NNNNNNNNNNNNNNNNNNNN | 1 |
| I | 41 | hCoV-19/USA/TX-HMH-3422/2020\|EPI_ISL_544180\|2020-0 | ···················· | ···················· | ···················· | ···················· | NNNNNNNNNNNNNNNNNNNN | NNNNNNNNNNNNNNNNNNNN | 1 |
| I | 51 | hCoV-19/USA/TX-HMH-3440/2020\|EPI_ISL_544195\|2020-0 | ···················· | ···················· | ···················· | ···················· | NNNNNNNNNNNNNNNNNNNN | NNNNNNNNNNNNNNNNNNNN | 1 |
| I | 54 | hCoV-19/USA/TX-HMH-3444/2020\|EPI_ISL_544199\|2020-0 | ···················· | ···················· | ···················· | ···················· | NNNNNNNNNNNNNNNNNNNN | NNNNNNNNNNNNNNNNNNNN | 1 |
| I | 63 | hCoV-19/USA/TX-HMH-2589/2020\|EPI_ISL_545412\|2020-0 | ···················· | ···················· | ···················· | ···················· | NNNNNNNNNNNNNNNNNNNN | NNNNNNNNNNNNNNNNNNNN | 1 |
| I | 80 | hCoV-19/USA/TX-HMH-2673/2020\|EPI_ISL_545467\|2020-0 | ···················· | ··············C····· | ···················· | ···················· | NN···NNN·····NN····· | NNN···NNNN···NN···N· | 1 |
| I | 101 | hCoV-19/USA/TX-HMH-2711/2020\|EPI_ISL_545495\|2020-0 | ···················· | ···················· | ···················· | ···················· | NNNNNNNNNNNNNNNNNNNN | NNNNNNNNNNNNNNNNNNNN | 1 |
| I | 102 | hCoV-19/USA/TX-HMH-2712/2020\|EPI_ISL_545496\|2020-0 | ···················· | ···················· | ···················· | ···················· | NNNNNNNNNNNNNNNNNNNN | NNNNNNNNNNNNNNNNNNNN | 1 |
| I | 115 | hCoV-19/USA/TX-HMH-7375/2020\|EPI_ISL_546715\|2020-0 | ···················· | ·············A······ | ···················· | ···················· | NNNNNNNNNNNNNNNNNNNN | NNNNNNNNNNNNNNNNNNNN | 1 |
| I | 116 | hCoV-19/USA/TX-HMH-7378/2020\|EPI_ISL_546717\|2020-0 | ···················· | ···················· | ···················· | ···················· | NNNNNNNNNNNNNNNNNNNN | NNNNNNNNNNNNNNNNNNNN | 1 |
| I | 122 | hCoV-19/USA/TX-HMH-7385/2020\|EPI_ISL_546724\|2020-0 | ···················· | ·············A······ | ···················· | ···················· | NNNNNNNNNNNNNNNNNNNN | NNNNNNNNNNNNNNNNNNNN | 1 |
| I | 123 | hCoV-19/USA/TX-HMH-7386/2020\|EPI_ISL_546725\|2020-0 | ···················· | N··················· | ···················· | ···················· | ··············NN···· | NN·····NN··········· | 1 |
| I | 124 | hCoV-19/USA/TX-HMH-7387/2020\|EPI_ISL_546726\|2020-0 | ···················· | ·············A······ | ···················· | ···················· | NNNNNNNNNNNNNNNNNNNN | NNNNNNNNNNNNNNNNNNNN | 1 |
| I | 132 | hCoV-19/USA/TX-HMH-7400/2020\|EPI_ISL_546739\|2020-0 | ···················· | ···················· | ···················· | ···················· | NNNNNNNNNNNNNNNNNNNN | NNNNNNNNNNNNNNNNNNNN | 1 |
| I | 138 | hCoV-19/USA/TX-HMH-7408/2020\|EPI_ISL_546745\|2020-0 | ···················· | ······A············· | ···················· | ···················· | NNNNNNNNNNNNNNNNNNNN | NNNNNNNNNNNNNNNNNNNN | 1 |
| I | 139 | hCoV-19/USA/TX-HMH-7409/2020\|EPI_ISL_546746\|2020-0 | ···················· | ···················· | ···················· | ···················· | NNNNNNNNNNNNNNNNNNNN | NNNNNNNNNNNNNNNNNNNN | 1 |
| I | 143 | hCoV-19/USA/TX-HMH-7416/2020\|EPI_ISL_546752\|2020-0 | ···················· | ·············A······ | ···················· | ···················· | NNNNNNNNNNNNNNNNNNNN | NNNNNNNNNNNNNNNNNNNN | 1 |
| I | 146 | hCoV-19/USA/TX-HMH-7421/2020\|EPI_ISL_546755\|2020-0 | ···················· | ···················· | ···················· | ···················· | NNNNNNNNNNNNNNNNNNNN | NNNNNNNNNNNNNNNNNNNN | 1 |
| I | 147 | hCoV-19/USA/TX-HMH-7423/2020\|EPI_ISL_546756\|2020-0 | ···················· | ···················· | ···················· | ···················· | NNNNNNNNNNNNNNNNNNNN | NNNNNNNNNNNNNNNNNNNN | 1 |
| I | 148 | hCoV-19/USA/TX-HMH-7428/2020\|EPI_ISL_546760\|2020-0 | ···················· | ·············A······ | ···················· | ···················· | NNNNNNNNNNNNNNNNNNNN | NNNNNNNNNNNNNNNNNNNN | 1 |
| I | 149 | hCoV-19/USA/TX-HMH-7431/2020\|EPI_ISL_546763\|2020-0 | ···················· | ···················· | ···················· | ···················· | NNNNNNNNNNNNNNNNNNNN | NNNNNNNNNNNNNNNNNNNN | 1 |
| I | 159 | hCoV-19/USA/TX-HMH-6061/2020\|EPI_ISL_547203\|2020-0 | ···················· | ···················· | ···················· | ···················· | NNNNNNNNNNNNNNNNNNNN | NNNNNNNNNNNNNNNNNNNN | 1 |
| I | 162 | hCoV-19/USA/TX-HMH-6064/2020\|EPI_ISL_547206\|2020-0 | ···················· | ···················· | ···················· | ···················· | NNNNNNNNNNNNNNNNNNNN | NNNNNNNNNNNNNNNNNNNN | 1 |
| I | 163 | hCoV-19/USA/TX-HMH-6065/2020\|EPI_ISL_547207\|2020-0 | ···················· | ·············A······ | ···················· | ···················· | NNNNNNNNNNNNNNNNNNNN | NNNNNNNNNNNNNNNNNNNN | 1 |
| I | 164 | hCoV-19/USA/TX-HMH-6066/2020\|EPI_ISL_547208\|2020-0 | ···················· | ···················· | ···················· | ···················· | NNNNNNNNNNNNNNNNNNNN | NNNNNNNNNNNNNNNNNNNN | 1 |
| I | 166 | hCoV-19/USA/TX-HMH-6068/2020\|EPI_ISL_547210\|2020-0 | ···················· | ···················· | ···················· | ···················· | NNNNNNNNNNNNNNNNNNNN | NNNNNNNNNNNNNNNNNNNN | 1 |
| I | 167 | hCoV-19/USA/TX-HMH-6070/2020\|EPI_ISL_547211\|2020-0 | ···················· | ···················· | ···················· | ···················· | NNNNNNNNNNNNNNNNNNNN | NNNNNNNNNNNNNNNNNNNN | 1 |
| I | 168 | hCoV-19/USA/TX-HMH-6071/2020\|EPI_ISL_547212\|2020-0 | ···················· | ···················· | ···················· | ···················· | NNNNNNNNNNNNNNNNNNNN | NNNNNNNNNNNNNNNNNNNN | 1 |
| I | 170 | hCoV-19/USA/TX-HMH-6075/2020\|EPI_ISL_547214\|2020-0 | ···················· | ···················· | ···················· | ···················· | NNNNNNNNNNNNNNNNNNNN | NNNNNNNNNNNNNNNNNNNN | 1 |
| I | 174 | hCoV-19/USA/TX-HMH-6080/2020\|EPI_ISL_547218\|2020-0 | ···················· | ·············A······ | ···················· | ···················· | NNNNNNNNNNNNNNNNNNNN | NNNNNNNNNNNNNNNNNNNN | 1 |
| I | 175 | hCoV-19/USA/TX-HMH-6081/2020\|EPI_ISL_547219\|2020-0 | ···················· | ···················· | ···················· | ···················· | NNNNNNNNNNNNNNNNNNNN | NNNNNNNNNNNNNNNNNNNN | 1 |
| I | 176 | hCoV-19/USA/TX-HMH-6083/2020\|EPI_ISL_547221\|2020-0 | ···················· | ···················· | ···················· | ···················· | NNNNNNNNNNNNNNNNNNNN | NNNNNNNNNNNNNNNNNNNN | 1 |
| I | 178 | hCoV-19/USA/TX-HMH-6085/2020\|EPI_ISL_547223\|2020-0 | ···················· | ···················· | ···················· | ···················· | NNNNNNNNNNNNNNNNNNNN | NNNNNNNNNNNNNNNNNNNN | 1 |
| I | 179 | hCoV-19/USA/TX-HMH-6086/2020\|EPI_ISL_547224\|2020-0 | ···················· | ···················· | ···················· | ···················· | N··N·······N··N····· | NNN····NN··········· | 1 |
| I | 180 | hCoV-19/USA/TX-HMH-6087/2020\|EPI_ISL_547225\|2020-0 | ···················· | ···················· | ···················· | ···················· | NNNNNNNNNNNNNNNNNNNN | NNNNNNNNNNNNNNNNNNNN | 1 |
| I | 181 | hCoV-19/USA/TX-HMH-6088/2020\|EPI_ISL_547226\|2020-0 | ···················· | ···················· | ···················· | ···················· | NNNNNNNNNNNNNNNNNNNN | NNNNNNNNNNNNNNNNNNNN | 1 |
| I | 183 | hCoV-19/USA/TX-HMH-6091/2020\|EPI_ISL_547230\|2020-0 | ···················· | ···················· | ···················· | ···················· | NNNNNNNNNNNNNNNNNNNN | NNNNNNNNNNNNNNNNNNNN | 1 |
| I | 184 | hCoV-19/USA/TX-HMH-6095/2020\|EPI_ISL_547233\|2020-0 | ···················· | ·············A······ | ···················· | ···················· | NNNNNNNNNNNNNNNNNNNN | NNNNNNNNNNNNNNNNNNNN | 1 |
| I | 185 | hCoV-19/USA/TX-HMH-6096/2020\|EPI_ISL_547234\|2020-0 | ···················· | ···················· | ···················· | ···················· | NNNNNNNNNNNNNNNNNNNN | NNNNNNNNNNNNNNNNNNNN | 1 |
| I | 186 | hCoV-19/USA/TX-HMH-6098/2020\|EPI_ISL_547235\|2020-0 | ···················· | ···················· | ···················· | ···················· | NNNNNNNNNNNNNNNNNNNN | NNNNNNNNNNNNNNNNNNNN | 1 |
| I | 189 | hCoV-19/USA/TX-HMH-6102/2020\|EPI_ISL_547238\|2020-0 | ···················· | ···················· | ···················· | ···················· | NNNNNNNNNNNNNNNNNNNN | NNNNNNNNNNNNNNNNNNNN | 1 |
| I | 190 | hCoV-19/USA/TX-HMH-6103/2020\|EPI_ISL_547239\|2020-0 | ···················· | ···················· | ···················· | ···················· | NNNNNNNNNNNNNNNNNNNN | NNNNNNNNNNNNNNNNNNNN | 1 |
| I | 281 | hCoV-19/USA/TX-HMH-3147/2020\|EPI_ISL_544026\|2020-0 | ···················· | ···················· | ···················· | ···················· | NNNNNNNNNNNNNNNNNNNN | NNNNNNNNNNNNNNNNNNNN | 1 |
| I | 282 | hCoV-19/USA/TX-HMH-3148/2020\|EPI_ISL_544027\|2020-0 | ···················· | ···················· | ···················· | ···················· | NNNNNNNNNNNNNNNNNNNN | NNNNNNNNNNNNNNNNNNNN | 1 |
| I | 302 | hCoV-19/USA/TX-HMH-3292/2020\|EPI_ISL_544109\|2020-0 | ···················· | ···················· | ···················· | ···················· | NNNNNNNNNNNNNNNNNNNN | NNNNNNNNNNNNNNNNNNNN | 1 |
| I | 312 | hCoV-19/USA/TX-HMH-3310/2020\|EPI_ISL_544122\|2020-0 | ···················· | ···················· | ···················· | ···················· | NNNNNNNNNNNNNNNNNNNN | NNNNNNNNNNNNNNNNNNNN | 1 |
| I | 355 | hCoV-19/USA/TX-HMH-2623/2020\|EPI_ISL_545433\|2020-0 | ···················· | ···················· | ···················· | ···················· | NNNNNNNNNNNNNNNNNNNN | NNNNNNNNNNNNNNNNNNNN | 1 |
| I | 375 | hCoV-19/USA/TX-HMH-2658/2020\|EPI_ISL_545456\|2020-0 | ···················· | ·A·················· | ···················· | ···················· | NNNNNNNNNNNNNNNNNNNN | NNNNNNNNNNNNNNNNNNNN | 1 |
| I | 382 | hCoV-19/USA/TX-HMH-7320/2020\|EPI_ISL_546674\|2020-0 | ···················· | ···················· | ···················· | ···················· | ···················· | NN·····NN··········· | 1 |
| I | 384 | hCoV-19/USA/TX-HMH-7327/2020\|EPI_ISL_546676\|2020-0 | ···················· | ···················· | ···················· | ···················· | NNNNNNNNNNNNNNNNNNNN | NNNNNNNNNNNNNNNNNNNN | 1 |
| I | 387 | hCoV-19/USA/TX-HMH-7330/2020\|EPI_ISL_546679\|2020-0 | ···················· | ···················· | ···················· | ···················· | NNNNNNNNNNNNNNNNNNNN | NNNNNNNNNNNNNNNNNNNN | 1 |
| I | 391 | hCoV-19/USA/TX-HMH-7337/2020\|EPI_ISL_546684\|2020-0 | ···················· | ···················· | ···················· | ···················· | NNNNNNNNNNNNNNNNNNNN | NNNNNNNNNNNNNNNNNNNN | 1 |
| I | 392 | hCoV-19/USA/TX-HMH-7339/2020\|EPI_ISL_546686\|2020-0 | ···················· | ···················· | ···················· | ···················· | NNNNNNNNNNNNNNNNNNNN | NNNNNNNNNNNNNNNNNNNN | 1 |
| I | 396 | hCoV-19/USA/TX-HMH-7343/2020\|EPI_ISL_546690\|2020-0 | ···················· | ···················· | ···················· | ···················· | NNNNNNNNNNNNNNNNNNNN | NNNNNNNNNNNNNNNNNNNN | 1 |
| I | 399 | hCoV-19/USA/TX-HMH-7348/2020\|EPI_ISL_546693\|2020-0 | ···················· | ···················· | ···················· | ···················· | NNNNNNNNNNNNNNNNNNNN | NNNNNNNNNNNNNNNNNNNN | 1 |
| I | 400 | hCoV-19/USA/TX-HMH-7349/2020\|EPI_ISL_546694\|2020-0 | ···················· | ···················· | ···················· | ···················· | ···N··········NNN··· | NNNN···N············ | 1 |
| I | 402 | hCoV-19/USA/TX-HMH-7354/2020\|EPI_ISL_546699\|2020-0 | ···················· | ···················· | ···················· | ···················· | NNNNNNNNNNNNNNNNNNNN | NNNNNNNNNNNNNNNNNNNN | 1 |
| I | 403 | hCoV-19/USA/TX-HMH-7424/2020\|EPI_ISL_546757\|2020-0 | ···················· | ···················· | ···················· | ···················· | NNNNNNNNNNNNNNNNNNNN | NNNNNNNNNNNNNNNNNNNN | 1 |
| I | 404 | hCoV-19/USA/TX-HMH-7426/2020\|EPI_ISL_546758\|2020-0 | ···················· | ···················· | ···················· | ···················· | NNNNNNNNNNNNNNNNNNNN | NNNNNNNNNNNNNNNNNNNN | 1 |
| I | 405 | hCoV-19/USA/TX-HMH-7427/2020\|EPI_ISL_546759\|2020-0 | ···················· | ···················· | ···················· | ···················· | ··············NN···· | NN········NN··N····· | 1 |
| I | 406 | hCoV-19/USA/TX-HMH-7432/2020\|EPI_ISL_546764\|2020-0 | ···················· | ···················· | ···················· | ···················· | NNNNNNNNNNNNNNNNNNNN | NNNNNNNNNNNNNNNNNNNN | 1 |
| I | 407 | hCoV-19/USA/TX-HMH-7433/2020\|EPI_ISL_546765\|2020-0 | ···················· | ···················· | ···················· | ···················· | NNNNNNNNNNNNNNNNNNNN | NNNNNNNNNNNNNNNNNNNN | 1 |
| I | 408 | hCoV-19/USA/TX-HMH-7435/2020\|EPI_ISL_546766\|2020-0 | ···················· | ···················· | ···················· | ···················· | NNNNNNNNNNNNNNNNNNNN | NNNNNNNNNNNNNNNNNNNN | 1 |
| I | 409 | hCoV-19/USA/TX-HMH-7439/2020\|EPI_ISL_546769\|2020-0 | ···················· | ·············A······ | ···················· | ···················· | NNNNNNNNNNNNNNNNNNNN | NNNNNNNNNNNNNNNNNNNN | 1 |
| I | 410 | hCoV-19/USA/TX-HMH-7440/2020\|EPI_ISL_546770\|2020-0 | ···················· | ·············A······ | ···················· | ···················· | NNNNNNNNNNNNNNNNNNNN | NNNNNNNNNNNNNNNNNNNN | 1 |
| I | 412 | hCoV-19/USA/TX-HMH-7448/2020\|EPI_ISL_546774\|2020-0 | ···················· | ···················· | ···················· | ···················· | NNNNNNNNNNNNNNNNNNNN | NNNNNNNNNNNNNNNNNNNN | 1 |
| I | 414 | hCoV-19/USA/TX-HMH-7450/2020\|EPI_ISL_546776\|2020-0 | ···················· | ·············A······ | ···················· | ···················· | NNNNNNNNNNNNNNNNNNNN | NNNNNNNNNNNNNNNNNNNN | 1 |
| I | 415 | hCoV-19/USA/TX-HMH-7451/2020\|EPI_ISL_546777\|2020-0 | ···················· | ·············A······ | ···················· | ···················· | ··N···N·N·····NN···· | NN·N···NNN···NN····· | 1 |
| I | 416 | hCoV-19/USA/TX-HMH-7452/2020\|EPI_ISL_546778\|2020-0 | ···················· | ···················· | ···················· | ···················· | NNNNNNNNNNNNNNNNNNNN | NNNNNNNNNNNNNNNNNNNN | 1 |
| I | 417 | hCoV-19/USA/TX-HMH-7454/2020\|EPI_ISL_546779\|2020-0 | ···················· | ···················· | ···················· | ···················· | NNNNNNNNNNNNNNNNNNNN | NNNNNNNNNNNNNNNNNNNN | 1 |
| I | 418 | hCoV-19/USA/TX-HMH-7459/2020\|EPI_ISL_546780\|2020-0 | ···················· | ···················· | ···················· | ···················· | NNNNNNNNNNNNNNNNNNNN | NNNNNNNNNNNNNNNNNNNN | 1 |
| I | 419 | hCoV-19/USA/TX-HMH-7460/2020\|EPI_ISL_546781\|2020-0 | ···················· | ···················· | ···················· | ···················· | NNNNNNNNNNNNNNNNNNNN | NNNNNNNNNNNNNNNNNNNN | 1 |
| I | 420 | hCoV-19/USA/TX-HMH-7462/2020\|EPI_ISL_546782\|2020-0 | ···················· | ···················· | ···················· | ···················· | NNNNNNNNNNNNNNNNNNNN | NNNNNNNNNNNNNNNNNNNN | 1 |

| | | | | | | | | | |
|---|---|---|---|---|---|---|---|---|---|
| I | 421 | hCoV-19/USA/TX-HMH-7465/2020\|EPI_ISL_546783\|2020-0 | ···················· | ···················· | ···················· | ···················· | NNNNNNNNNNNNNNNNNNNN | NNNNNNNNNNNNNNNNNNNN | 1 |
| I | 422 | hCoV-19/USA/TX-HMH-7467/2020\|EPI_ISL_546785\|2020-0 | ···················· | ···················· | ···················· | ···················· | NNNNNNNNNNNNNNNNNNNN | NNNNNNNNNNNNNNNNNNNN | 1 |
| I | 424 | hCoV-19/USA/TX-HMH-7470/2020\|EPI_ISL_546787\|2020-0 | ···················· | ···················· | ···················· | ···················· | NNNNNNNNNNNNNNNNNNNN | NNNNNNNNNNNNNNNNNNNN | 1 |
| I | 425 | hCoV-19/USA/TX-HMH-7473/2020\|EPI_ISL_546789\|2020-0 | ···················· | ···················· | ···················· | ···················· | NNNNNNNNNNNNNNNNNNNN | NNNNNNNNNNNNNNNNNNNN | 1 |
| I | 426 | hCoV-19/USA/TX-HMH-7493/2020\|EPI_ISL_546800\|2020-0 | ···················· | ···················· | ···················· | ···················· | NNNNNNNNNNNNNNNNNNNN | NNNNNNNNNNNNNNNNNNNN | 1 |
| I | 427 | hCoV-19/USA/TX-HMH-7495/2020\|EPI_ISL_546801\|2020-0 | ···················· | ···················· | ···················· | ···················· | NNNNNNNNNNNNNNNNNNNN | NNNNNNNNNNNNNNNNNNNN | 1 |
| I | 428 | hCoV-19/USA/TX-HMH-7496/2020\|EPI_ISL_546802\|2020-0 | ···················· | ···················· | ···················· | ···················· | NNNNNNNNNNNNNNNNNNNN | NNNNNNNNNNNNNNNNNNNN | 1 |
| I | 429 | hCoV-19/USA/TX-HMH-7500/2020\|EPI_ISL_546803\|2020-0 | ···················· | ···················· | ···················· | ···················· | NNNNNNNNNNNNNNNNNNNN | NNNNNNNNNNNNNNNNNNNN | 1 |
| I | 430 | hCoV-19/USA/TX-HMH-7501/2020\|EPI_ISL_546804\|2020-0 | ···················· | ···················· | ···················· | ···················· | NNNNNNNNNNNNNNNNNNNN | NNNNNNNNNNNNNNNNNNNN | 1 |
| I | 431 | hCoV-19/USA/TX-HMH-7503/2020\|EPI_ISL_546805\|2020-0 | ···················· | ···················· | ···················· | ···················· | NN············NNN··· | NNNN···NN·NN·NN····· | 1 |
| I | 432 | hCoV-19/USA/TX-HMH-7506/2020\|EPI_ISL_546807\|2020-0 | ···················· | ···················· | ···················· | ···················· | NNNNNNNNNNNNNNNNNNNN | NNNNNNNNNNNNNNNNNNNN | 1 |
| I | 436 | hCoV-19/USA/TX-HMH-7512/2020\|EPI_ISL_546812\|2020-0 | ···················· | ···················· | ···················· | ···················· | NNNNNNNNNNNNNNNNNNNN | NNNNNNNNNNNNNNNNNNNN | 1 |
| I | 440 | hCoV-19/USA/TX-HMH-7525/2020\|EPI_ISL_546820\|2020-0 | ···················· | ···················· | ···················· | ···················· | ···················· | NN·····N········N··· | 1 |
| I | 441 | hCoV-19/USA/TX-HMH-6104/2020\|EPI_ISL_547240\|2020-0 | ···················· | ·A·················· | ···················· | ···················· | NNNNNNNNNNNNNNNNNNNN | NNNNNNNNNNNNNNNNNNNN | 1 |
| I | 442 | hCoV-19/USA/TX-HMH-6109/2020\|EPI_ISL_547244\|2020-0 | ···················· | ···················· | ···················· | ···················· | NNNNNNNNNNNNNNNNNNNN | NNNNNNNNNNNNNNNNNNNN | 1 |
| I | 444 | hCoV-19/USA/TX-HMH-6110/2020\|EPI_ISL_547246\|2020-0 | ···················· | ···················· | ···················· | ···················· | NNNNNNNNNNNNNNNNNNNN | NNNNNNNNNNNNNNNNNNNN | 1 |
| I | 445 | hCoV-19/USA/TX-HMH-6112/2020\|EPI_ISL_547248\|2020-0 | ···················· | ···················· | ···················· | ···················· | NNNNNNNNNNNNNNNNNNNN | NNNNNNNNNNNNNNNNNNNN | 1 |
| I | 446 | hCoV-19/USA/TX-HMH-6114/2020\|EPI_ISL_547250\|2020-0 | ···················· | ·············A······ | ···················· | ···················· | NNNNNNNNNNNNNNNNNNNN | NNNNNNNNNNNNNNNNNNNN | 1 |
| I | 447 | hCoV-19/USA/TX-HMH-6115/2020\|EPI_ISL_547251\|2020-0 | ···················· | ···················· | ···················· | ···················· | NNNNNNNNNNNNNNNNNNNN | NNNNNNNNNNNNNNNNNNNN | 1 |
| I | 448 | hCoV-19/USA/TX-HMH-6116/2020\|EPI_ISL_547252\|2020-0 | ···················· | ···················· | ···················· | ···················· | NNNNNNNNNNNNNNNNNNNN | NNNNNNNNNNNNNNNNNNNN | 1 |
| I | 449 | hCoV-19/USA/TX-HMH-6118/2020\|EPI_ISL_547253\|2020-0 | ···················· | ·············A······ | ···················· | ···················· | NNNNNNNNNNNNNNNNNNNN | NNNNNNNNNNNNNNNNNNNN | 1 |
| I | 450 | hCoV-19/USA/TX-HMH-6119/2020\|EPI_ISL_547254\|2020-0 | ···················· | ·············A······ | ···················· | ···················· | NNNNNNNNNNNNNNNNNNNN | NNNNNNNNNNNNNNNNNNNN | 1 |
| I | 452 | hCoV-19/USA/TX-HMH-6121/2020\|EPI_ISL_547256\|2020-0 | ···················· | ···················· | ···················· | ···················· | NNNNNNNNNNNNNNNNNNNN | NNNNNNNNNNNNNNNNNNNN | 1 |
| I | 453 | hCoV-19/USA/TX-HMH-6126/2020\|EPI_ISL_547259\|2020-0 | ···················· | ·············A······ | ···················· | ···················· | NNNNNNNNNNNNNNNNNNNN | NNNNNNNNNNNNNNNNNNNN | 1 |
| I | 454 | hCoV-19/USA/TX-HMH-6128/2020\|EPI_ISL_547261\|2020-0 | ···················· | ···················· | ···················· | ···················· | ··············N····· | NN·N···NN·NN·NN····· | 1 |
| I | 456 | hCoV-19/USA/TX-HMH-6132/2020\|EPI_ISL_547263\|2020-0 | ···················· | ···················· | ···················· | ···················· | NNNNNNNNNNNNNNNNNNNN | NNNNNNNNNNNNNNNNNNNN | 1 |
| I | 457 | hCoV-19/USA/TX-HMH-6133/2020\|EPI_ISL_547264\|2020-0 | ···················· | ···················· | ···················· | ···················· | ···N·NNNN·····N····· | NNNN···NN····NNN···· | 1 |
| I | 458 | hCoV-19/USA/TX-HMH-6139/2020\|EPI_ISL_547266\|2020-0 | ···················· | ···················· | ···················· | ···················· | NNNNNNNNNNNNNNNNNNNN | NNNNNNNNNNNNNNNNNNNN | 1 |
| I | 459 | hCoV-19/USA/TX-HMH-6145/2020\|EPI_ISL_547268\|2020-0 | ···················· | ···················· | ···················· | ···················· | NNNNNNNNNNNNNNNNNNNN | NNNNNNNNNNNNNNNNNNNN | 1 |
| I | 461 | hCoV-19/USA/TX-HMH-6148/2020\|EPI_ISL_547270\|2020-0 | ···················· | ·············A······ | ···················· | ···················· | NNNNNNNNNNNNNNNNNNNN | NNNNNNNNNNNNNNNNNNNN | 1 |
| I | 462 | hCoV-19/USA/TX-HMH-6149/2020\|EPI_ISL_547271\|2020-0 | ···················· | ···················· | ···················· | ···················· | NNNNNNNNNNNNNNNNNNNN | NNNNNNNNNNNNNNNNNNNN | 1 |
| I | 463 | hCoV-19/USA/TX-HMH-615/2020\|EPI_ISL_547272\|2020-04 | ···················· | ···················· | ···················· | ···················· | NNNNNNNNNNNNNNNNNNNN | ···················· | 1 |
| I | 464 | hCoV-19/USA/TX-HMH-6150/2020\|EPI_ISL_547273\|2020-0 | ···················· | ···················· | ···················· | ···················· | NNNNNNNNNNNNNNNNNNNN | NNNNNNNNNNNNNNNNNNNN | 1 |
| I | 465 | hCoV-19/USA/TX-HMH-6151/2020\|EPI_ISL_547274\|2020-0 | ···················· | ···················· | ···················· | ···················· | NNNNNNNNNNNNNNNNNNNN | NNNNNNNNNNNNNNNNNNNN | 1 |
| I | 466 | hCoV-19/USA/TX-HMH-6153/2020\|EPI_ISL_547276\|2020-0 | ···················· | ···················· | ···················· | ···················· | NNNNNNNNNNNNNNNNNNNN | NNNNNNNNNNNNNNNNNNNN | 1 |
| I | 467 | hCoV-19/USA/TX-HMH-6156/2020\|EPI_ISL_547277\|2020-0 | ···················· | ···················· | ···················· | ···················· | NNNNNNNNNNNNNNNNNNNN | NNNNNNNNNNNNNNNNNNNN | 1 |
| I | 468 | hCoV-19/USA/TX-HMH-6157/2020\|EPI_ISL_547278\|2020-0 | ···················· | ···················· | ···················· | ···················· | NNNNNNNNNNNNNNNNNNNN | NNNNNNNNNNNNNNNNNNNN | 1 |
| I | 469 | hCoV-19/USA/TX-HMH-6158/2020\|EPI_ISL_547279\|2020-0 | ···················· | ···················· | ···················· | ···················· | NNNNNNNNNNNNNNNNNNNN | NNNNNNNNNNNNNNNNNNNN | 1 |
| I | 476 | hCoV-19/USA/TX-HMH-6173/2020\|EPI_ISL_547291\|2020-0 | ···················· | ···················· | ···················· | ···················· | NNNNNNNNNNNNNNNNNNNN | NNNNNNNNNNNNNNNNNNNN | 1 |
| I | 514 | hCoV-19/USA/TX-HMH-3113/2020\|EPI_ISL_544001\|2020-0 | ···················· | ···················· | ···················· | ···················· | NNNNNNNNNNNNNNNNNNNN | NNNNNNNNNNNNNNNNNNNN | 1 |
| I | 547 | hCoV-19/USA/TX-HMH-3208/2020\|EPI_ISL_544077\|2020-0 | ···················· | ···················· | ···················· | ···················· | NNNNNNNNNNNNNNNNNNNN | NNNNNNNNNNNNNNNNNNNN | 1 |
| I | 563 | hCoV-19/USA/TX-HMH-5599/2020\|EPI_ISL_544891\|2020-0 | ···················· | ···················· | ···················· | ···················· | NNNNNNNNNNNNNNNNNNNN | NNNNNNNNNNNNNNNNNNNN | 1 |
| I | 565 | hCoV-19/USA/TX-HMH-5604/2020\|EPI_ISL_544895\|2020-0 | ···················· | ···················· | ···················· | ···················· | NNNNNNNNNNNNNNNNNNNN | NNNNNNNNNNNNNNNNNNNN | 1 |
| I | 573 | hCoV-19/USA/TX-HMH-5643/2020\|EPI_ISL_544929\|2020-0 | ···················· | NNNNNNNNNNNNNNNNNNNN | ···················· | ···················· | ···················· | ···················· | 1 |
| I | 574 | hCoV-19/USA/TX-HMH-5644/2020\|EPI_ISL_544930\|2020-0 | ···················· | ···················· | ···················· | ···················· | ···N··········N····· | NN·····NN········N·· | 1 |
| I | 581 | hCoV-19/USA/TX-HMH-5657/2020\|EPI_ISL_544941\|2020-0 | ···················· | ···················· | ···················· | ···················· | NNNNNNNNNNNNNNNNNNNN | NNNNNNNNNNNNNNNNNNNN | 1 |
| I | 582 | hCoV-19/USA/TX-HMH-5662/2020\|EPI_ISL_544944\|2020-0 | ···················· | ···················· | ···················· | ···················· | NNNNNNNNNNNNNNNNNNNN | NNNNNNNNNNNNNNNNNNNN | 1 |
| I | 583 | hCoV-19/USA/TX-HMH-5663/2020\|EPI_ISL_544945\|2020-0 | ···················· | ···················· | ···················· | ···················· | NNNNNNNNNNNNNNNNNNNN | NNNNNNNNNNNNNNNNNNNN | 1 |
| I | 584 | hCoV-19/USA/TX-HMH-5664/2020\|EPI_ISL_544946\|2020-0 | ···················· | ···················· | ···················· | ···················· | NNNNNNNNNNNNNNNNNNNN | NNNNNNNNNNNNNNNNNNNN | 1 |
| I | 585 | hCoV-19/USA/TX-HMH-5665/2020\|EPI_ISL_544947\|2020-0 | ···················· | ···················· | ···················· | ···················· | ··············N····· | NN·N···NN··········· | 1 |
| I | 588 | hCoV-19/USA/TX-HMH-5673/2020\|EPI_ISL_544953\|2020-0 | ···················· | ···················· | ···················· | ···················· | N·······N·NNNNN·N··· | ·NNN···NN·NN·NN····· | 1 |
| I | 613 | hCoV-19/USA/TX-HMH-2436/2020\|EPI_ISL_545308\|2020-0 | ···················· | ···················· | ···················· | ···················· | ··············N····· | NN·····NN··········· | 1 |
| I | 661 | hCoV-19/USA/TX-HMH-7209/2020\|EPI_ISL_546600\|2020-0 | ···················· | ···················· | ···················· | ···················· | NNNNNNNNNNNNNNNNNNNN | NNNNNNNNNNNNNNNNNNNN | 1 |
| I | 662 | hCoV-19/USA/TX-HMH-7211/2020\|EPI_ISL_546601\|2020-0 | ···················· | ···················· | ···················· | ···················· | NNNNNNNNNNNNNNNNNNNN | NNNNNNNNNNNNNNNNNNNN | 1 |
| I | 665 | hCoV-19/USA/TX-HMH-7216/2020\|EPI_ISL_546605\|2020-0 | ···················· | ···················· | ···················· | ···················· | NNNNNNNNNNNNNNNNNNNN | NNNNNNNNNNNNNNNNNNNN | 1 |
| I | 666 | hCoV-19/USA/TX-HMH-7217/2020\|EPI_ISL_546606\|2020-0 | ···················· | ···················· | ···················· | ···················· | NNNNNNNNNNNNNNNNNNNN | NNNNNNNNNNNNNNNNNNNN | 1 |
| I | 669 | hCoV-19/USA/TX-HMH-7226/2020\|EPI_ISL_546611\|2020-0 | ···················· | ·············A······ | ···················· | ···················· | ···N················ | NN·N····N··········· | 1 |
| I | 675 | hCoV-19/USA/TX-HMH-7236/2020\|EPI_ISL_546618\|2020-0 | ···················· | ··········T········· | ···················· | ···················· | NNNNNNNNNNNNNNNNNNNN | NNNNNNNNNNNNNNNNNNNN | 1 |
| I | 680 | hCoV-19/USA/TX-HMH-7250/2020\|EPI_ISL_546626\|2020-0 | ···················· | ·············A······ | ···················· | ···················· | NNNNNNNNNNNNNNNNNNNN | NNNNNNNNNNNNNNNNNNNN | 1 |
| I | 681 | hCoV-19/USA/TX-HMH-7252/2020\|EPI_ISL_546627\|2020-0 | ···················· | ···················· | ···················· | ···················· | NNNNNNNNNNNNNNNNNNNN | NNNNNNNNNNNNNNNNNNNN | 1 |
| I | 682 | hCoV-19/USA/TX-HMH-7256/2020\|EPI_ISL_546629\|2020-0 | ···················· | ···················· | ···················· | ···················· | ··············NN···· | NN·····NN··········· | 1 |
| I | 686 | hCoV-19/USA/TX-HMH-7269/2020\|EPI_ISL_546638\|2020-0 | ···················· | ···················· | ···················· | ···················· | NNNNNNNNNNNNNNNNNNNN | NNNNNNNNNNNNNNNNNNNN | 1 |
| I | 688 | hCoV-19/USA/TX-HMH-7271/2020\|EPI_ISL_546640\|2020-0 | ···················· | ···················· | ···················· | ···················· | NNNNNNNNNNNNNNNNNNNN | NNNNNNNNNNNNNNNNNNNN | 1 |
| I | 689 | hCoV-19/USA/TX-HMH-7272/2020\|EPI_ISL_546641\|2020-0 | ···················· | ···················· | ···················· | ···················· | NNNNNNNNNNNNNNNNNNNN | NNNNNNNNNNNNNNNNNNNN | 1 |
| I | 690 | hCoV-19/USA/TX-HMH-7273/2020\|EPI_ISL_546642\|2020-0 | ···················· | ···················· | ···················· | ···················· | NNNNNNNNNNNNNNNNNNNN | NNNNNNNNNNNNNNNNNNNN | 1 |
| I | 691 | hCoV-19/USA/TX-HMH-7277/2020\|EPI_ISL_546645\|2020-0 | ···················· | ···················· | ···················· | ···················· | NNNNNNNNNNNNNNNNNNNN | NNNNNNNNNNNNNNNNNNNN | 1 |
| I | 694 | hCoV-19/USA/TX-HMH-7294/2020\|EPI_ISL_546651\|2020-0 | ···················· | ···················· | ···················· | ···················· | NNNNNNNNNNNNNNNNNNNN | NNNNNNNNNNNNNNNNNNNN | 1 |
| I | 699 | hCoV-19/USA/TX-HMH-7301/2020\|EPI_ISL_546658\|2020-0 | ···················· | ···················· | ···················· | ···················· | NNNNNNNNNNNNNNNNNNNN | NNNNNNNNNNNNNNNNNNNN | 1 |
| I | 702 | hCoV-19/USA/TX-HMH-7306/2020\|EPI_ISL_546663\|2020-0 | ···················· | ···················· | ···················· | ···················· | NNNNNNNNNNNNNNNNNNNN | NNNNNNNNNNNNNNNNNNNN | 1 |
| I | 703 | hCoV-19/USA/TX-HMH-7307/2020\|EPI_ISL_546664\|2020-0 | ···················· | ···················· | ···················· | ···················· | NNNNNNNNNNNNNNNNNNNN | NNNNNNNNNNNNNNNNNNNN | 1 |
| I | 705 | hCoV-19/USA/TX-HMH-7311/2020\|EPI_ISL_546668\|2020-0 | ···················· | ···················· | ···················· | ···················· | NNNNNNNNNNNNNNNNNNNN | NNNNNNNNNNNNNNNNNNNN | 1 |
| I | 707 | hCoV-19/USA/TX-HMH-5908/2020\|EPI_ISL_547101\|2020-0 | ···················· | ···················· | ···················· | ···················· | NNNNNNNNNNNNNNNNNNNN | NNNNNNNNNNNNNNNNNNNN | 1 |
| I | 708 | hCoV-19/USA/TX-HMH-5910/2020\|EPI_ISL_547102\|2020-0 | ···················· | ···················· | ···················· | ···················· | NNNNNNNNNNNNNNNNNNNN | NNNNNNNNNNNNNNNNNNNN | 1 |
| I | 710 | hCoV-19/USA/TX-HMH-5913/2020\|EPI_ISL_547105\|2020-0 | ···················· | ···················· | ···················· | ···················· | NNNNNNNNNNNNNNNNNNNN | NNNNNNNNNNNNNNNNNNNN | 1 |
| I | 714 | hCoV-19/USA/TX-HMH-5943/2020\|EPI_ISL_547110\|2020-0 | ···················· | ······A············· | ···················· | ···················· | NNNNNNNNNNNNNNNNNNNN | NNNNNNNNNNNNNNNNNNNN | 1 |
| I | 716 | hCoV-19/USA/TX-HMH-5945/2020\|EPI_ISL_547112\|2020-0 | ···················· | ···················· | ···················· | ···················· | NNNNNNNNNNNNNNNNNNNN | NNNNNNNNNNNNNNNNNNNN | 1 |
| I | 717 | hCoV-19/USA/TX-HMH-5946/2020\|EPI_ISL_547113\|2020-0 | ···················· | ·············A······ | ···················· | ···················· | NNNNNNNNNNNNNNNNNNNN | NNNNNNNNNNNNNNNNNNNN | 1 |
| I | 752 | hCoV-19/USA/CA-CZB-3686/2020\|EPI_ISL_548414\|2020-0 | ···················· | ···················· | ···················· | ···················· | NNNNNNNNNNNNNNNNNNNN | ···················· | 1 |
| I | 753 | hCoV-19/USA/CA-CZB-3706/2020\|EPI_ISL_548417\|2020-0 | ···················· | ·A·················· | ···················· | ···················· | NNNNNNNNNNNNNNNNNNNN | ···················· | 1 |
| I | 754 | hCoV-19/USA/CA-CZB-3718/2020\|EPI_ISL_548420\|2020-0 | ···················· | ·A·················· | ···················· | ···················· | NNNNNNNNNNNNNNNNNNNN | NNNNNNNNNNNNNNNNNNNN | 1 |

| | | | | | | | | | |
|---|---|---|---|---|---|---|---|---|---|
| I | 755 | hCoV-19/USA/CA-CZB-3699/2020\|EPI_ISL_548423\|2020-0 | ···················· | ···················· | ···················· | ···················· | NNNNNNNNNNNNNNNNNNNN | NNNNNNNNNNNNNNN····N | 1 |
| I | 756 | hCoV-19/USA/CA-CZB-3721/2020\|EPI_ISL_548425\|2020-0 | ···················· | ···················· | ···················· | ···················· | NNNNNNNNNNNNNNNNNNNN | NNNNNNNNNNNNNNNNNNNN | 1 |
| I | 757 | hCoV-19/USA/CA-CZB-3694/2020\|EPI_ISL_548426\|2020-0 | ···················· | ···················· | ···················· | ···················· | NNNNNNNNNNNNNNNNNNNN | NNNNNNNNNNNNNNNNNNNN | 1 |
| I | 758 | hCoV-19/USA/CA-CZB-3836/2020\|EPI_ISL_548428\|2020-0 | ···················· | ·A·················· | ···················· | ···················· | NNNNNNNNNNNNNNNNNNNN | NNNNNNNNNNNNNNNNNNNN | 1 |
| I | 759 | hCoV-19/USA/CA-CZB-3690/2020\|EPI_ISL_548429\|2020-0 | ···················· | ···················· | ···················· | ···················· | NNNNNNNNNNNNNNNNNNNN | NNNNNNNNNNNNNNNNNNNN | 1 |
| I | 761 | hCoV-19/USA/CA-CZB-3744/2020\|EPI_ISL_548431\|2020-0 | ···················· | ···················· | ···················· | ···················· | NNNNNNNNNNNNNNNNNNNN | ···················· | 1 |
| I | 762 | hCoV-19/USA/CA-CZB-3701/2020\|EPI_ISL_548433\|2020-0 | ···················· | ·A·················· | ···················· | ···················· | NNNNNNNNNNNNNNNNNNNN | NNNNNNNNNNNNNNNNNNNN | 1 |
| I | 763 | hCoV-19/USA/CA-CZB-3717/2020\|EPI_ISL_548434\|2020-0 | ···················· | ·A·················· | ···················· | ···················· | NNNNNNNNNNNNNNNNNNNN | NNNNNNNNNNNNNNNNNNNN | 1 |
| I | 767 | hCoV-19/USA/CA-CZB-3726/2020\|EPI_ISL_548444\|2020-0 | ···················· | ···················· | ···················· | ···················· | NNNNNNNNNNN········· | ···················· | 1 |
| I | 778 | hCoV-19/USA/TX-HMH-5609/2020\|EPI_ISL_544900\|2020-0 | ···················· | ···················· | ···················· | ···················· | NNNNNNNNNNNNNNNNNNNN | NNNNNNNNNNNNNNNNNNNN | 1 |
| I | 779 | hCoV-19/USA/TX-HMH-5611/2020\|EPI_ISL_544902\|2020-0 | ···················· | ···················· | ···················· | ···················· | NNNNNNNNNNNNNNNNNNNN | NNNNNNNNNNNNNNNNNNNN | 1 |
| I | 786 | hCoV-19/USA/TX-HMH-5624/2020\|EPI_ISL_544911\|2020-0 | ···················· | ···················· | ···················· | ···················· | NNNNNNNNNNNNNNNNNNNN | NNNNNNNNNNNNNNNNNNNN | 1 |
| I | 876 | hCoV-19/USA/TX-HMH-7152/2020\|EPI_ISL_546549\|2020-0 | ···················· | ···················· | ···················· | ···················· | NNNNNNNNNNNNNNNNNNNN | NNNNNNNNNNNNNNNNNNNN | 1 |
| I | 877 | hCoV-19/USA/TX-HMH-7153/2020\|EPI_ISL_546550\|2020-0 | ···················· | ···················· | ···················· | ···················· | NNNNNNNNNNNNNNNNNNNN | NNNNNNNNNNNNNNNNNNNN | 1 |
| I | 878 | hCoV-19/USA/TX-HMH-7155/2020\|EPI_ISL_546551\|2020-0 | ···················· | ···················· | ···················· | ···················· | NNNNNNNNNNNNNNNNNNNN | NNNNNNNNNNNNNNNNNNNN | 1 |
| I | 879 | hCoV-19/USA/TX-HMH-7157/2020\|EPI_ISL_546552\|2020-0 | ···················· | ···················· | ···················· | ···················· | NNNNNNNNNNNNNNNNNNNN | NNNNNNNNNNNNNNNNNNNN | 1 |
| I | 880 | hCoV-19/USA/TX-HMH-7159/2020\|EPI_ISL_546554\|2020-0 | ···················· | ···················· | ···················· | ···················· | NNNNNNNNNNNNNNNNNNNN | NNNNNNNNNNNNNNNNNNNN | 1 |
| I | 884 | hCoV-19/USA/TX-HMH-7162/2020\|EPI_ISL_546558\|2020-0 | ···················· | ···················· | ···················· | ···················· | NNNNNNNNNNNNNNNNNNNN | NNNNNNNNNNNNNNNNNNNN | 1 |
| I | 890 | hCoV-19/USA/TX-HMH-7172/2020\|EPI_ISL_546568\|2020-0 | ···················· | ···················· | ···················· | ···················· | NNNNNNNNNNNNNNNNNNNN | NNNNNNNNNNNNNNNNNNNN | 1 |
| I | 894 | hCoV-19/USA/TX-HMH-7176/2020\|EPI_ISL_546572\|2020-0 | ···················· | ···················· | ···················· | ···················· | NNNNNNNNNNNNNNNNNNNN | NNNNNNNNNNNNNNNNNNNN | 1 |
| I | 897 | hCoV-19/USA/TX-HMH-7183/2020\|EPI_ISL_546579\|2020-0 | ···················· | ·A·················· | ···················· | ···················· | NNNNNNNNNNNNNNNNNNNN | NNNNNNNNNNNNNNNNNNNN | 1 |
| I | 899 | hCoV-19/USA/TX-HMH-7186/2020\|EPI_ISL_546582\|2020-0 | ···················· | ···················· | ···················· | ···················· | NNNNNNNNNNNNNNNNNNNN | NNNNNNNNNNNNNNNNNNNN | 1 |
| I | 900 | hCoV-19/USA/TX-HMH-7190/2020\|EPI_ISL_546584\|2020-0 | ···················· | ···················· | ···················· | ···················· | NNNNNNNNNNNNNNNNNNNN | NNNNNNNNNNNNNNNNNNNN | 1 |
| I | 903 | hCoV-19/USA/TX-HMH-7193/2020\|EPI_ISL_546587\|2020-0 | ···················· | ···················· | ···················· | ···················· | NNNNNNNNNNNNNNNNNNNN | NNNNNNNNNNNNNNNNNNNN | 1 |
| I | 904 | hCoV-19/USA/TX-HMH-7198/2020\|EPI_ISL_546592\|2020-0 | ···················· | ···················· | ···················· | ···················· | ··············NN···· | NNN····N············ | 1 |
| I | 910 | hCoV-19/USA/TX-HMH-7356/2020\|EPI_ISL_546701\|2020-0 | ···················· | ···················· | ···················· | ···················· | NNNNNNNNNNNNNNNNNNNN | NNNNNNNNNNNNNNNNNNNN | 1 |
| I | 911 | hCoV-19/USA/TX-HMH-5957/2020\|EPI_ISL_547121\|2020-0 | ···················· | ···················· | ···················· | ···················· | NNNNNNNNNNNNNNNNNNNN | NNNNNNNNNNNNNNNNNNNN | 1 |
| I | 915 | hCoV-19/USA/TX-HMH-5961/2020\|EPI_ISL_547125\|2020-0 | ···················· | ·············A······ | ···················· | ···················· | ··NN··········NNN··· | NN·····N···N·NNN···· | 1 |
| I | 917 | hCoV-19/USA/TX-HMH-5963/2020\|EPI_ISL_547127\|2020-0 | ···················· | ···················· | ···················· | ···················· | NNNNNNNNNNNNNNNNNNNN | NNNNNNNNNNNNNNNNNNNN | 1 |
| I | 918 | hCoV-19/USA/TX-HMH-5965/2020\|EPI_ISL_547129\|2020-0 | ···················· | ···················· | ···················· | ···················· | NNNNNNNNNNNNNNNNNNNN | NNNNNNNNNNNNNNNNNNNN | 1 |
| I | 919 | hCoV-19/USA/TX-HMH-5966/2020\|EPI_ISL_547130\|2020-0 | ···················· | ···················· | ···················· | ···················· | NNNNNNNNNNNNNNNNNNNN | NNNNNNNNNNNNNNNNNNNN | 1 |
| I | 920 | hCoV-19/USA/TX-HMH-5968/2020\|EPI_ISL_547131\|2020-0 | ···················· | ·A·················· | ···················· | ···················· | NNNNNNNNNNNNNNNNNNNN | NNNNNNNNNNNNNNNNNNNN | 1 |
| I | 922 | hCoV-19/USA/TX-HMH-5970/2020\|EPI_ISL_547133\|2020-0 | ···················· | ···················· | ···················· | ···················· | NNNNNNNNNNNNNNNNNNNN | NNNNNNNNNNNNNNNNNNNN | 1 |
| I | 924 | hCoV-19/USA/TX-HMH-5976/2020\|EPI_ISL_547138\|2020-0 | ···················· | ···················· | ···················· | ···················· | ·N·N····N····NNN·NN· | NN·N···NN····NN····· | 1 |
| I | 927 | hCoV-19/USA/TX-HMH-5983/2020\|EPI_ISL_547143\|2020-0 | ···················· | ···················· | ···················· | ···················· | NNNNNNNNNNNNNNNNNNNN | NNNNNNNNNNNNNNNNNNNN | 1 |
| I | 930 | hCoV-19/USA/TX-HMH-5988/2020\|EPI_ISL_547148\|2020-0 | ···················· | ···················· | ···················· | ···················· | NNNNNNNNNNNNNNNNNNNN | NNNNNNNNNNNNNNNNNNNN | 1 |
| I | 931 | hCoV-19/USA/TX-HMH-5989/2020\|EPI_ISL_547149\|2020-0 | ···················· | ···················· | ···················· | ···················· | NNNNNNNNNNNNNNNNNNNN | NNNNNNNNNNNNNNNNNNNN | 1 |
| I | 938 | hCoV-19/USA/TX-HMH-6005/2020\|EPI_ISL_547161\|2020-0 | ···················· | ···················· | ···················· | ···················· | ···N····N·····NN·N·· | NNNN·NNN············ | 1 |
| I | 946 | hCoV-19/USA/TX-HMH-6019/2020\|EPI_ISL_547171\|2020-0 | ···················· | ···················· | ···················· | ···················· | NNNNNNNNNNNNNNNNNNNN | NNNNNNNNNNNNNNNNNNNN | 1 |
| I | 949 | hCoV-19/USA/TX-HMH-6022/2020\|EPI_ISL_547174\|2020-0 | ···················· | ···T················ | ···················· | ···················· | NNNNNNNNNNNNNNNNNNNN | NNNNNNNNNNNNNNNNNNNN | 1 |
| I | 959 | hCoV-19/USA/TX-HMH-6040/2020\|EPI_ISL_547185\|2020-0 | ···················· | ···················· | ···················· | ···················· | NNNNNNNNNNNNNNNNNNNN | NN·NNN·NNNN··NNN···· | 1 |
| I | 960 | hCoV-19/USA/TX-HMH-6041/2020\|EPI_ISL_547186\|2020-0 | ···················· | ···················· | ···················· | ···················· | NNNNNNNNNNNNNNNNNNNN | NNNNNNNNNNNNNNNNNNNN | 1 |
| I | 961 | hCoV-19/USA/TX-HMH-6042/2020\|EPI_ISL_547187\|2020-0 | ···················· | ···················· | ···················· | ···················· | NNNNNNNNNNNNNNNNNNNN | NNNNNNNNNNNNNNNNNNNN | 1 |
| I | 965 | hCoV-19/USA/TX-HMH-6051/2020\|EPI_ISL_547194\|2020-0 | ···················· | ·····A·············· | ···················· | ···················· | NNNNNNNNNNNNNNNNNNNN | NNNNNNNNNNNNNNNNNNNN | 1 |
| I | 967 | hCoV-19/USA/TX-HMH-6054/2020\|EPI_ISL_547197\|2020-0 | ···················· | ·····A·············· | ···················· | ···················· | NNNNNNNNNNNNNNNNNNNN | NNNNNNNNNNNNNNNNNNNN | 1 |
| I | 1007 | hCoV-19/USA/CA-CZB-3703/2020\|EPI_ISL_548482\|2020-0 | ···················· | ·A·················· | ···················· | ···················· | NNNNNNNNNNN········· | ···················· | 1 |
| I | 1028 | hCoV-19/USA/TX-HMH-5432/2020\|EPI_ISL_544770\|2020-0 | ···················· | ···················· | ···················· | ···················· | NNNNNNNNNNNNNNNNNNNN | NNNNNNNNNNNNNNNNNNNN | 1 |
| I | 1032 | hCoV-19/USA/TX-HMH-5438/2020\|EPI_ISL_544776\|2020-0 | ···················· | ·············A······ | ···················· | ···················· | NNNNNNNNNNNNNNNNNNNN | NNNNNNNNNNNNNNNNNNNN | 1 |
| I | 1033 | hCoV-19/USA/TX-HMH-5440/2020\|EPI_ISL_544777\|2020-0 | ···················· | ···················· | ···················· | ···················· | NNNNNNNNNNNNNNNNNNNN | NNNNNNNNNNNNNNNNNNNN | 1 |
| I | 1034 | hCoV-19/USA/TX-HMH-5441/2020\|EPI_ISL_544778\|2020-0 | ···················· | ·A·················· | ···················· | ···················· | NNNNNNNNNNNNNNNNNNNN | NNNNNNNNNNNNNNNNNNNN | 1 |
| I | 1035 | hCoV-19/USA/TX-HMH-5442/2020\|EPI_ISL_544779\|2020-0 | ···················· | ···················· | ···················· | ···················· | NNNNNNNNNNNNNNNNNNNN | NNNNNNNNNNNNNNNNNNNN | 1 |
| I | 1036 | hCoV-19/USA/TX-HMH-5443/2020\|EPI_ISL_544780\|2020-0 | ···················· | ···················· | ···················· | ···················· | NNNNNNNNNNNNNNNNNNNN | NNNNNNNNNNNNNNNNNNNN | 1 |
| I | 1038 | hCoV-19/USA/TX-HMH-5447/2020\|EPI_ISL_544782\|2020-0 | ···················· | ···················· | ···················· | ···················· | ·NNN····N·NNN·NN·N·· | NNNN···NN·NN········ | 1 |
| I | 1039 | hCoV-19/USA/TX-HMH-5448/2020\|EPI_ISL_544783\|2020-0 | ···················· | ·····A·············· | ···················· | ···················· | NNNNNNNNNNNNNNNNNNNN | NNNNNNNNNNNNNNNNNNNN | 1 |
| I | 1041 | hCoV-19/USA/TX-HMH-5455/2020\|EPI_ISL_544788\|2020-0 | ···················· | ···················· | ···················· | ···················· | NNNNNNNNNNNNNNNNNNNN | NNNNNNNNNNNNNNNNNNNN | 1 |
| I | 1042 | hCoV-19/USA/TX-HMH-5458/2020\|EPI_ISL_544791\|2020-0 | ···················· | ···················· | ···················· | ···················· | NNNNNNNNNNNNNNNNNNNN | NNNNNNNNNNNNNNNNNNNN | 1 |
| I | 1043 | hCoV-19/USA/TX-HMH-5459/2020\|EPI_ISL_544792\|2020-0 | ···················· | ···················· | ···················· | ···················· | NNNNNNNNNNNNNNNNNNNN | NNNNNNNNNNNNNNNNNNNN | 1 |
| I | 1050 | hCoV-19/USA/TX-HMH-5477/2020\|EPI_ISL_544802\|2020-0 | ···················· | ···················· | ···················· | ···················· | ··············N···NN | NN·N·NNNN···NNN····· | 1 |
| I | 1051 | hCoV-19/USA/TX-HMH-5478/2020\|EPI_ISL_544803\|2020-0 | ···················· | ···················· | ···················· | ···················· | ·······N······N····N | NNNN···NN·NN·N····N· | 1 |
| I | 1056 | hCoV-19/USA/TX-HMH-5490/2020\|EPI_ISL_544813\|2020-0 | ···················· | ···················· | ···················· | ···················· | NNNNNNNNNNNNNNNNNNNN | NNNNNNNNNNNNNNNNNNNN | 1 |
| I | 1058 | hCoV-19/USA/TX-HMH-5493/2020\|EPI_ISL_544816\|2020-0 | ···················· | ···················· | ···················· | ···················· | NNNNNNNNNNNNNNNNNNNN | NNNNNNNNNNNNNNNNNNNN | 1 |
| I | 1059 | hCoV-19/USA/TX-HMH-5496/2020\|EPI_ISL_544819\|2020-0 | ···················· | ···················· | ···················· | ···················· | NNNNNNNNNNNNNNNNNNNN | NNNNNNNNNNNNNNNNNNNN | 1 |
| I | 1061 | hCoV-19/USA/TX-HMH-5498/2020\|EPI_ISL_544821\|2020-0 | ···················· | ·A·················· | ···················· | ···················· | NNNNNNNNNNNNNNNNNNNN | NNNNNNNNNNNNNNNNNNNN | 1 |
| I | 1063 | hCoV-19/USA/TX-HMH-550/2020\|EPI_ISL_544823\|2020-04 | ···················· | ···················· | ···················· | ···················· | NNNNNNNNNNNNNNNNNNNN | NNNNNNNNNNNNNNNNNNNN | 1 |
| I | 1064 | hCoV-19/USA/TX-HMH-5500/2020\|EPI_ISL_544824\|2020-0 | ···················· | ···················· | ···················· | ···················· | NNNNNNNNNNNNNNNNNNNN | NNNNNNNNNNNNNNNNNNNN | 1 |
| I | 1065 | hCoV-19/USA/TX-HMH-5501/2020\|EPI_ISL_544825\|2020-0 | ···················· | ···················· | ···················· | ···················· | NNNNNNNNNNNNNNNNNNNN | NNNNNNNNNNNNNNNNNNNN | 1 |
| I | 1066 | hCoV-19/USA/TX-HMH-5502/2020\|EPI_ISL_544826\|2020-0 | ···················· | ···················· | ···················· | ···················· | NNNNNNNNNNNNNNNNNNNN | NNNNNNNNNNNNNNNNNNNN | 1 |
| I | 1067 | hCoV-19/USA/TX-HMH-5504/2020\|EPI_ISL_544828\|2020-0 | ···················· | ···················· | ···················· | ···················· | NNNNNNNNNNNNNNNNNNNN | NNNNNNNNNNNNNNNNNNNN | 1 |
| I | 1069 | hCoV-19/USA/TX-HMH-5508/2020\|EPI_ISL_544831\|2020-0 | ···················· | ···················· | ···················· | ···················· | NNNNNNNNNNNNNNNNNNNN | NNNNNNNNNNNNNNNNNNNN | 1 |
| I | 1070 | hCoV-19/USA/TX-HMH-5509/2020\|EPI_ISL_544832\|2020-0 | ···················· | ·A·················· | ···················· | ···················· | NNNNNNNNNNNNNNNNNNNN | NNNNNNNNNNNNNNNNNNNN | 1 |
| I | 1071 | hCoV-19/USA/TX-HMH-551/2020\|EPI_ISL_544833\|2020-04 | ···················· | ···················· | ···················· | ···················· | NNNNNNNNNNN········· | ···················· | 1 |
| I | 1072 | hCoV-19/USA/TX-HMH-5510/2020\|EPI_ISL_544834\|2020-0 | ···················· | ·············A······ | ···················· | ···················· | NNNNNNNNNNNNNNNNNNNN | NNNNNNNNNNNNNNNNNNNN | 1 |
| I | 1078 | hCoV-19/USA/TX-HMH-5521/2020\|EPI_ISL_544842\|2020-0 | ···················· | ···················· | ···················· | ···················· | NNNNNNNNNNNNNNNNNNNN | NNNNNNNNNNNNNNNNNNNN | 1 |
| I | 1079 | hCoV-19/USA/TX-HMH-5523/2020\|EPI_ISL_544843\|2020-0 | ···················· | ···················· | ···················· | ···················· | NNNNNNNNNNNNNNNNNNNN | NNNNNNNNNNNNNNNNNNNN | 1 |
| I | 1080 | hCoV-19/USA/TX-HMH-5532/2020\|EPI_ISL_544844\|2020-0 | ···················· | ···················· | ···················· | ···················· | NNNNNNNNNNNNNNNNNNNN | NNNNNNNNNNNNNNNNNNNN | 1 |
| I | 1081 | hCoV-19/USA/TX-HMH-5535/2020\|EPI_ISL_544845\|2020-0 | ···················· | ···················· | ···················· | ···················· | NNNNNNNNNNNNNNNNNNNN | NNNNNNNNNNNNNNNNNNNN | 1 |
| I | 1082 | hCoV-19/USA/TX-HMH-5537/2020\|EPI_ISL_544847\|2020-0 | ···················· | ···················· | ···················· | ···················· | NNNNNNNNNNNNNNNNNNNN | NNNNNNNNNNNNNNNNNNNN | 1 |
| I | 1084 | hCoV-19/USA/TX-HMH-5539/2020\|EPI_ISL_544849\|2020-0 | ···················· | ···················· | ···················· | ···················· | NNNNNNNNNNNNNNNNNNNN | NNNNNNNNNNNNNNNNNNNN | 1 |
| I | 1086 | hCoV-19/USA/TX-HMH-5542/2020\|EPI_ISL_544851\|2020-0 | ···················· | ···················· | ···················· | ···················· | N·NNN·NN······NN···· | NNNN···············N | 1 |
| I | 1089 | hCoV-19/USA/TX-HMH-5547/2020\|EPI_ISL_544854\|2020-0 | ···················· | ·············A······ | ···················· | ···················· | NNNNNNNNNNNNNNNNNNNN | NNNNNNNNNNNNNNNNNNNN | 1 |

I 1090 hCoV-19/USA/TX-HMH-5548/2020|EPI_ISL_544855|2020-0 ···················· ···················· ···················· ···················· NNNNNNNNNNNNNNNNNNNN NNNNNNNNNNNNNNNNNNNN 1
I 1091 hCoV-19/USA/TX-HMH-5550/2020|EPI_ISL_544857|2020-0 ···················· ·············A······ ···················· ···················· NNNNNNNNNNNNNNNNNNNN NNNNNNNNNNNNNNNNNNNN 1
I 1092 hCoV-19/USA/TX-HMH-5557/2020|EPI_ISL_544859|2020-0 ···················· ···················· ···················· ···················· NNNNNNNNNNNNNNNNNNNN NNNNNNNNNNNNNNNNNNNN 1
I 1094 hCoV-19/USA/TX-HMH-5563/2020|EPI_ISL_544864|2020-0 ···················· ·············A······ ···················· ···················· NNNNNNNNNNNNNNNNNNNN NNNNNNNNNNNNNNNNNNNN 1
I 1095 hCoV-19/USA/TX-HMH-5565/2020|EPI_ISL_544866|2020-0 ···················· ···················· ···················· ···················· NNNNNNNNNNNNNNNNNNNN NNNNNNNNNNNNNNNNNNNN 1
I 1098 hCoV-19/USA/TX-HMH-5572/2020|EPI_ISL_544870|2020-0 ···················· ···················· ···················· ···················· NNNNNNNNNNNNNNNNNNNN NNNNNNNNNNNNNNNNNNNN 1
I 1099 hCoV-19/USA/TX-HMH-5573/2020|EPI_ISL_544871|2020-0 ···················· ·············A······ ···················· ···················· NNNNNNNNNNNNNNNNNNNN NNNNNNNNNNNNNNNNNNNN 1
I 1100 hCoV-19/USA/TX-HMH-5574/2020|EPI_ISL_544872|2020-0 ···················· ···················· ···················· ···················· NNNNNNNNNNNNNNNNNNNN NNNNNNNNNNNNNNNNNNNN 1
I 1102 hCoV-19/USA/TX-HMH-5576/2020|EPI_ISL_544874|2020-0 ···················· ···················· ···················· ···················· NNNNNNNNNNNNNNNNNNNN NNNNNNNNNNNNNNNNNNNN 1
I 1105 hCoV-19/USA/TX-HMH-5581/2020|EPI_ISL_544878|2020-0 ···················· ···················· ···················· ···················· NNNNNNNNNNNNNNNNNNNN NNNNNNNNNNNNNNNNNNNN 1
I 1110 hCoV-19/USA/TX-HMH-559/2020|EPI_ISL_544885|2020-04 ···················· ···················· ···················· ···················· NNNNNNNNNNNNNNNNNNNN ···················· 1
I 1112 hCoV-19/USA/TX-HMH-2243/2020|EPI_ISL_545200|2020-0 ···················· ···················· ···················· ···················· NNNNNNNNNNNNNNNNNNNN NNNNNNNNNNNNNNNNNNNN 1
I 1145 hCoV-19/USA/TX-HMH-7120/2020|EPI_ISL_546521|2020-0 ···················· ···················· ···················· ···················· NNNNNNNNNNNNNNNNNNNN NNNNNNNNNNNNNNNNNNNN 1
I 1147 hCoV-19/USA/TX-HMH-7122/2020|EPI_ISL_546523|2020-0 ···················· ···················· ···················· ···················· NNNNNNNNNNNNNNNNNNNN NNNNNNNNNNNNNNNNNNNN 1
I 1154 hCoV-19/USA/TX-HMH-7133/2020|EPI_ISL_546533|2020-0 ···················· ···················· ···················· ···················· NNNNNNNNNNNNNNNNNNNN NNNNNNNNNNNNNNNNNNNN 1
I 1155 hCoV-19/USA/TX-HMH-7136/2020|EPI_ISL_546536|2020-0 ···················· ···················· ···················· ···················· NNNNNNNNNNNNNNNNNNNN NNNNNNNNNNNNNNNNNNNN 1
I 1156 hCoV-19/USA/TX-HMH-7137/2020|EPI_ISL_546537|2020-0 ···················· ···················· ···················· ···················· NNNNNNNNNNNNNNNNNNNN NNNNNNNNNNNNNNNNNNNN 1
I 1158 hCoV-19/USA/TX-HMH-7140/2020|EPI_ISL_546539|2020-0 ···················· ···················· ···················· ···················· NNNNNNNNNNNNNNNNNNNN NNNNNNNNNNNNNNNNNNNN 1
I 1160 hCoV-19/USA/TX-HMH-7144/2020|EPI_ISL_546542|2020-0 ···················· ···················· ···················· ···················· NNNNNNNNNNNNNNNNNNNN NNNNNNNNNNNNNNNNNNNN 1
I 1161 hCoV-19/USA/TX-HMH-7147/2020|EPI_ISL_546545|2020-0 ···················· ···················· ···················· ···················· NNNNNNNNNNNNNNNNNNNN NNNNNNNNNNNNNNNNNNNN 1
I 1162 hCoV-19/USA/TX-HMH-7148/2020|EPI_ISL_546546|2020-0 ···················· ···················· ···················· ···················· NNNNNNNNNNNNNNNNNNNN NNNNNNNNNNNNNNNNNNNN 1
I 1174 hCoV-19/USA/TX-HMH-5785/2020|EPI_ISL_547012|2020-0 ···················· ···················· ···················· ···················· ·N·N·······N·NN··N·· NNNN·NNNN·NN········ 1
I 1179 hCoV-19/USA/TX-HMH-5790/2020|EPI_ISL_547017|2020-0 ···················· ···················· ···················· ···················· NNNNNNNNNNNNNNNNNNNN NNNNNNNNNNNNNNNNNNNN 1
I 1180 hCoV-19/USA/TX-HMH-5877/2020|EPI_ISL_547080|2020-0 ···················· ···················· ···················· ···················· NNNNNNNNNNNNNNNNNNNN NNNNNNNNNNNNNNNNNNNN 1
I 1181 hCoV-19/USA/TX-HMH-5878/2020|EPI_ISL_547081|2020-0 ···················· ···················· ···················· ···················· ··············N··N·N NNNN···N············ 1
I 1182 hCoV-19/USA/TX-HMH-5879/2020|EPI_ISL_547082|2020-0 ···················· ·············A······ ···················· ···················· NNNNNNNNNNNNNNNNNNNN NNNNNNNNNNNNNNNNNNNN 1
I 1184 hCoV-19/USA/TX-HMH-5882/2020|EPI_ISL_547085|2020-0 ···················· ···················· ···················· ···················· NNNNNNNNNNNNNNNNNNNN NNNNNNNNNNNNNNNNNNNN 1
I 1185 hCoV-19/USA/TX-HMH-5885/2020|EPI_ISL_547086|2020-0 ···················· ···················· ···················· ···················· NNNNNNNNNNNNNNNNNNNN NNNNNNNNNNNNNNNNNNNN 1
I 1188 hCoV-19/USA/TX-HMH-5895/2020|EPI_ISL_547091|2020-0 ···················· ···················· ···················· ···················· NNNNNNNNNNNNNNNNNNNN NNNNNNNNNNNNNNNNNNNN 1
I 1190 hCoV-19/USA/TX-HMH-590/2020|EPI_ISL_547094|2020-04 ···················· ···················· ···················· ···················· NNNNNNNNNNNNNNNNNN·· ···················· 1
I 1194 hCoV-19/USA/TX-HMH-5905/2020|EPI_ISL_547099|2020-0 ···················· ·············A······ ···················· ···················· NNNNNNNNNNNNNNNNNNNN NNNNNNNNNNNNNNNNNNNN 1
I 1284 hCoV-19/USA/TX-HMH-5283/2020|EPI_ISL_544652|2020-0 ···················· ···················· ···················· ···················· N···NNN·······NNN··· NN·····N············ 1
I 1285 hCoV-19/USA/TX-HMH-5284/2020|EPI_ISL_544653|2020-0 ···················· ···················· ···················· ···················· NNNNNNNNN····NN····N NNNN···NN·NNNNNN··N· 1
I 1286 hCoV-19/USA/TX-HMH-2066/2020|EPI_ISL_545100|2020-0 ···················· ···················· ···················· ···················· NNNNNNNNNNNNNNNNNNNN NNNNNNNNNNNNNNNNNNNN 1
I 1345 hCoV-19/USA/TX-HMH-2183/2020|EPI_ISL_545168|2020-0 ···················· ···················· ···················· ···················· NNNNNNNNNNNNNNNNNNNN NNNNNNNNNNNNNNNNNNNN 1
I 1351 hCoV-19/USA/TX-HMH-2197/2020|EPI_ISL_545175|2020-0 ···················· ···················· ···················· ···················· ··NNNNNN······N···NN NN·N···NNNNNNNN···NN 1
I 1356 hCoV-19/USA/TX-HMH-6405/2020|EPI_ISL_545976|2020-0 ···················· ·············A······ ···················· ···················· NNNNNNNNNNNNNNNNNNNN NNNNNNNNNNNNNNNNNNNN 1
I 1362 hCoV-19/USA/TX-HMH-6420/2020|EPI_ISL_545986|2020-0 ···················· ···················· ···················· ···················· NNNNNNNNNNNNNNNNNNNN NNNNNNNNNNNNNNNNNNNN 1
I 1363 hCoV-19/USA/TX-HMH-7008/2020|EPI_ISL_546427|2020-0 ···················· ···················· ···················· ···················· NNNNNNNNNNNNNNNNNNNN NNNNNNNNNNNNNNNNNNNN 1
I 1364 hCoV-19/USA/TX-HMH-7011/2020|EPI_ISL_546430|2020-0 ···················· ···················· ···················· ···················· NNNNNNNNNNNNNNNNNNNN NNNNNNNNNNNNNNNNNNNN 1
I 1365 hCoV-19/USA/TX-HMH-7013/2020|EPI_ISL_546432|2020-0 ···················· ···················· ···················· ···················· NNNNNNNNNNNNNNNNNNNN NNNNNNNNNNNNNNNNNNNN 1
I 1366 hCoV-19/USA/TX-HMH-7014/2020|EPI_ISL_546433|2020-0 ···················· ···················· ···················· ···················· NNNNNNNNNNNNNNNNNNNN NNNNNNNNNNNNNNNNNNNN 1
I 1367 hCoV-19/USA/TX-HMH-7015/2020|EPI_ISL_546434|2020-0 ···················· ···················· ···················· ···················· NNNNNNNNNNNNNNNNNNNN NNNNNNNNNNNNNNNNNNNN 1
I 1371 hCoV-19/USA/TX-HMH-702/2020|EPI_ISL_546439|2020-04 ···················· ···················· ···················· ···················· NNNN················ ···················· 1
I 1375 hCoV-19/USA/TX-HMH-7027/2020|EPI_ISL_546446|2020-0 ···················· ···················· ···················· ···················· NNNNNNNNNNNNNNNNNNNN NNNNNNNNNNNNNNNNNNNN 1
I 1378 hCoV-19/USA/TX-HMH-7031/2020|EPI_ISL_546449|2020-0 ···················· ···················· ···················· ···················· NNNNNNNNNNNNNNNNNNNN NNNNNNNNNNNNNNNNNNNN 1
I 1383 hCoV-19/USA/TX-HMH-704/2020|EPI_ISL_546457|2020-04 ···················· ······R············· ···················· ···················· NNNNNNNNNNNNNNNNNNNN ···················· 1
I 1390 hCoV-19/USA/TX-HMH-706/2020|EPI_ISL_546471|2020-04 ···················· ···················· ···················· ···················· NNNNNNNNNNNNNNNNNNNN ···················· 1
I 1391 hCoV-19/USA/TX-HMH-7062/2020|EPI_ISL_546473|2020-0 ···················· ···················· ···················· ···················· NNNNNNNNNNNNNNNNNNNN NNNNNNNNNNNNNNNNNNNN 1
I 1406 hCoV-19/USA/TX-HMH-7085/2020|EPI_ISL_546492|2020-0 ···················· ·············A······ ···················· ···················· NNNNNNNNNNNNNNNNNNNN NNNNNNNNNNNNNNNNNNNN 1
I 1414 hCoV-19/USA/TX-HMH-5799/2020|EPI_ISL_547023|2020-0 ···················· ·············A······ ···················· ···················· NNNNNNNNNNNNNNNNNNNN NNNNNNNNNNNNNNNNNNNN 1
I 1415 hCoV-19/USA/TX-HMH-5803/2020|EPI_ISL_547025|2020-0 ···················· ···················· ···················· ···················· NNNNNNNNNNNNNNNNNNNN NNNNNNNNNNNNNNNNNNNN 1
I 1422 hCoV-19/USA/TX-HMH-5814/2020|EPI_ISL_547036|2020-0 ···················· ·············A······ ···················· ···················· NNNNNNNNNNNNNNNNNNNN NNNNNNNNNNNNNNNNNNNN 1
I 1425 hCoV-19/USA/TX-HMH-5821/2020|EPI_ISL_547041|2020-0 ···················· ···················· ···················· ···················· NNNNNNNNNNNNNNNNNNNN NNNNNNNNNNNNNNNNNNNN 1
I 1426 hCoV-19/USA/TX-HMH-5823/2020|EPI_ISL_547042|2020-0 ···················· ···················· ···················· ···················· NNNNNNNNNNNNNNNNNNNN NNNNNNNNNNNNNNNNNNNN 1
I 1427 hCoV-19/USA/TX-HMH-5825/2020|EPI_ISL_547044|2020-0 ···················· ···················· ···················· ···················· NNNNNNNNNNNNNNNNNNNN NNNNNNNNNNNNNNNNNNNN 1
I 1428 hCoV-19/USA/TX-HMH-5829/2020|EPI_ISL_547045|2020-0 ···················· ···················· ···················· ···················· NNNNNNNNNNNNNNNNNNNN NNNNNNNNNNNNNNNNNNNN 1
I 1430 hCoV-19/USA/TX-HMH-5831/2020|EPI_ISL_547047|2020-0 ···················· ···················· ···················· ···················· NNNNNNNNNNNNNNNNNNNN NNNNNNNNNNNNNNNNNNNN 1
I 1432 hCoV-19/USA/TX-HMH-5834/2020|EPI_ISL_547049|2020-0 ···················· ···················· ···················· ···················· NNNNNNNNNNNNNNNNNNNN NNNNNNNNNNNNNNNNNNNN 1
I 1433 hCoV-19/USA/TX-HMH-5836/2020|EPI_ISL_547050|2020-0 ···················· ···················· ···················· ···················· ········N··········· NN·N···NN··········· 1
I 1434 hCoV-19/USA/TX-HMH-5837/2020|EPI_ISL_547051|2020-0 ···················· ···················· ···················· ···················· NNNNNNNNNNNNNNNNNNNN NNNNNNNNNNNNNNNNNNNN 1
I 1435 hCoV-19/USA/TX-HMH-5840/2020|EPI_ISL_547054|2020-0 ···················· ···················· ···················· ···················· NNNNNNNNNNNNNNNNNNNN NNNNNNNNNNNNNNNNNNNN 1
I 1437 hCoV-19/USA/TX-HMH-5843/2020|EPI_ISL_547057|2020-0 ···················· ···················· ···················· ···················· NNNNNNNNNNNNNNNNNNNN NNNNNNNNNNNNNNNNNNNN 1
I 1438 hCoV-19/USA/TX-HMH-5849/2020|EPI_ISL_547060|2020-0 ···················· ···················· ···················· ···················· NNNNNNNNNNNNNNNNNNNN NNNNNNNNNNNNNNNNNNNN 1
I 1440 hCoV-19/USA/TX-HMH-5854/2020|EPI_ISL_547064|2020-0 ···················· ···················· ···················· ···················· NNNNNNNNNNNNNNNNNNNN NNNNNNNNNNNNNNNNNNNN 1
I 1441 hCoV-19/USA/TX-HMH-5856/2020|EPI_ISL_547066|2020-0 ···················· ···················· ···················· ···················· NNNNNNNNNNNNNNNNNNNN NNNNNNNNNNNNNNNNNNNN 1
I 1442 hCoV-19/USA/TX-HMH-5857/2020|EPI_ISL_547067|2020-0 ···················· ···················· ···················· ···················· ···N·NNN····NNNN···· NN·N················ 1
I 1443 hCoV-19/USA/TX-HMH-5858/2020|EPI_ISL_547068|2020-0 ···················· ···················· ···················· ···················· NNNNNNNNNNNNNNNNNNNN NNNNNNNNNNNNNNNNNNNN 1
I 1447 hCoV-19/USA/TX-HMH-5864/2020|EPI_ISL_547073|2020-0 ···················· ·············A······ ···················· ···················· NNNNNNNNNNNNNNNNNNNN NNNNNNNNNNNNNNNNNNNN 1
I 1534 hCoV-19/USA/CA-CZB-3714/2020|EPI_ISL_548394|2020-0 ···················· ·A·················· ···················· ···················· NNNNNNNNNNNNNNN····· ···················· 1
I 1535 hCoV-19/USA/CA-CZB-3708/2020|EPI_ISL_548395|2020-0 ···················· ···················· ···················· ···················· NNNNNNNNNNNNNNNNNNNN NNNNNNNNNNNNNNNNNNNN 1
I 1538 hCoV-19/USA/TX-HMH-5279/2020|EPI_ISL_544648|2020-0 ···················· ···················· ···················· ···················· NNNNNNNNNNNNNNNNNNNN NNNNNNNNNNNNNNNNNNNN 1
I 1545 hCoV-19/USA/TX-HMH-530/2020|EPI_ISL_544662|2020-04 ···················· ···················· ···················· ···················· NNNNNNNNNNNNNNNNNNNN NNNNNNNNNNNNNNNNNNNN 1
I 1547 hCoV-19/USA/TX-HMH-5304/2020|EPI_ISL_544665|2020-0 ···················· ···················· ···················· ···················· ······NN······N····N NN·N···NN·NN·N······ 1
I 1549 hCoV-19/USA/TX-HMH-5306/2020|EPI_ISL_544667|2020-0 ···················· ···················· ···················· ···················· NNNNNNNNNNNNNNNNNNNN NNNNNNNNNNNNNNNNNNNN 1
I 1550 hCoV-19/USA/TX-HMH-5307/2020|EPI_ISL_544668|2020-0 ···················· ·············A······ ···················· ···················· NNNNNNNNNNNNNNNNNNNN NNNNNNNNNNNNNNNNNNNN 1
I 1551 hCoV-19/USA/TX-HMH-5308/2020|EPI_ISL_544669|2020-0 ···················· ···················· ···················· ···················· NNNNNNNNNNNNNNNNNNNN NNNNNNNNNNNNNNNNNNNN 1
I 1552 hCoV-19/USA/TX-HMH-5309/2020|EPI_ISL_544670|2020-0 ···················· ···················· ···················· ···················· NNNNNNNNNNNNNNNNNNNN NNNNNNNNNNNNNNNNNNNN 1
I 1553 hCoV-19/USA/TX-HMH-5311/2020|EPI_ISL_544672|2020-0 ···················· ···················· ···················· ···················· NNNNNNNNNNN·NNN··N·· NNN····NNN···NNN··NN 1
I 1558 hCoV-19/USA/TX-HMH-5322/2020|EPI_ISL_544679|2020-0 ···················· ···················· ···················· ···················· NNNNNNNNNNNNNNNNNNNN NNNNNNNNNNNNNNNNNNNN 1

| | | | | | | | | | |
|---|---|---|---|---|---|---|---|---|---|
| I | 1565 | hCoV-19/USA/TX-HMH-5330/2020\|EPI_ISL_544688\|2020-0 | ···················· | ···················· | ···················· | ···················· | NNNNNNNNNNNNNNNNNNNN | NNNNNNNNNNNNNNNNNNNN | 1 |
| I | 1567 | hCoV-19/USA/TX-HMH-5334/2020\|EPI_ISL_544691\|2020-0 | ···················· | ···················· | ···················· | ···················· | NNNNNNNNNNNNNNNNNNNN | NNNNNNNNNNNNNNNNNNNN | 1 |
| I | 1568 | hCoV-19/USA/TX-HMH-5336/2020\|EPI_ISL_544692\|2020-0 | ···················· | ···················· | ···················· | ···················· | NNNNNNNNNNNNNNNNNNNN | NNNNNNNNNNNNNNNNNNNN | 1 |
| I | 1569 | hCoV-19/USA/TX-HMH-5337/2020\|EPI_ISL_544693\|2020-0 | ···················· | ·········A·········· | ···················· | ···················· | NNNNNNNNNNNNNNNNNNNN | NNNNNNNNNNNNNNNNNNNN | 1 |
| I | 1572 | hCoV-19/USA/TX-HMH-534/2020\|EPI_ISL_544696\|2020-04 | ···················· | ···················· | ···················· | ···················· | NNNNNNNNNNNNNNNNNNNN | NNNNNNNNNNNNNNNNNNNN | 1 |
| I | 1575 | hCoV-19/USA/TX-HMH-5343/2020\|EPI_ISL_544699\|2020-0 | ···················· | ···················· | ···················· | ···················· | NNNNNNNNNNNNNNNNNNNN | NNNNNNNNNNNNNNNNNNNN | 1 |
| I | 1577 | hCoV-19/USA/TX-HMH-5345/2020\|EPI_ISL_544701\|2020-0 | ···················· | ···················· | ···················· | ···················· | NNNNNNNNNNNNNNNNNNNN | NNNNNNNNNNNNNNNNNNNN | 1 |
| I | 1584 | hCoV-19/USA/TX-HMH-5353/2020\|EPI_ISL_544709\|2020-0 | ···················· | ···················· | ···················· | ···················· | NNNNNNNNNNNNNNNNNNNN | NNNNNNNNNNNNNNNNNNNN | 1 |
| I | 1586 | hCoV-19/USA/TX-HMH-5357/2020\|EPI_ISL_544712\|2020-0 | ···················· | ···················· | ···················· | ···················· | NNNNNNNNNNNNNNNNNNNN | NNNNNNNNNNNNNNNNNNNN | 1 |
| I | 1591 | hCoV-19/USA/TX-HMH-5363/2020\|EPI_ISL_544717\|2020-0 | ···················· | ···················· | ···················· | ···················· | NNNNNNNNNNNNNNNNNNNN | NNNNNNNNNNNNNNNNNNNN | 1 |
| I | 1592 | hCoV-19/USA/TX-HMH-5364/2020\|EPI_ISL_544718\|2020-0 | ···················· | ···················· | ···················· | ···················· | NNNNNNNNNNNNNNNNNNNN | NNNNNNNNNNNNNNNNNNNN | 1 |
| I | 1593 | hCoV-19/USA/TX-HMH-5365/2020\|EPI_ISL_544719\|2020-0 | ···················· | ···················· | ···················· | ···················· | NNNNNNNNNNNNNNNNNNNN | NNNNNNNNNNNNNNNNNNNN | 1 |
| I | 1602 | hCoV-19/USA/TX-HMH-5376/2020\|EPI_ISL_544728\|2020-0 | ···················· | ···················· | ···················· | ···················· | NNNNNNNNNNNNNNNNNNNN | NNNNNNNNNNNNNNNNNNNN | 1 |
| I | 1611 | hCoV-19/USA/TX-HMH-5397/2020\|EPI_ISL_544743\|2020-0 | ···················· | ···················· | ···················· | ···················· | ·N···NNN····N·NN·NNN | NNN····NNNNN····N··· | 1 |
| I | 1612 | hCoV-19/USA/TX-HMH-5399/2020\|EPI_ISL_544744\|2020-0 | ···················· | ···················· | ···················· | ···················· | NNNNNNNNNNNNNNNNNNNN | NNNNNNNNNNNNNNNNNNNN | 1 |
| I | 1613 | hCoV-19/USA/TX-HMH-5402/2020\|EPI_ISL_544747\|2020-0 | ···················· | ···················· | ···················· | ···················· | NNNNNNNNNNNNNNNNNNNN | NNNNNNNNNNNNNNNNNNNN | 1 |
| I | 1615 | hCoV-19/USA/TX-HMH-5407/2020\|EPI_ISL_544752\|2020-0 | ···················· | ············A······· | ···················· | ···················· | NNNNNNNNNNNNNNNNNNNN | NNNNNNNNNNNNNNNNNNNN | 1 |
| I | 1617 | hCoV-19/USA/TX-HMH-5413/2020\|EPI_ISL_544757\|2020-0 | ···················· | ···················· | ···················· | ···················· | ···················· | NN····NN············ | 1 |
| I | 1621 | hCoV-19/USA/TX-HMH-5419/2020\|EPI_ISL_544763\|2020-0 | ···················· | ···················· | ···················· | ···················· | ·N·N··········NN···· | NNNN··NNNNNN·····NNN | 1 |
| I | 1623 | hCoV-19/USA/TX-HMH-5421/2020\|EPI_ISL_544765\|2020-0 | ···················· | ···················· | ···················· | ···················· | NNNNNNNNNNNNNNNNNNNN | NNNNNNNNNNNNNNNNNNNN | 1 |
| I | 1645 | hCoV-19/USA/TX-HMH-6433/2020\|EPI_ISL_545992\|2020-0 | ···················· | ···················· | ···················· | ···················· | ·············NNN···· | NNNN················ | 1 |
| I | 1652 | hCoV-19/USA/TX-HMH-6978/2020\|EPI_ISL_546403\|2020-0 | ···················· | ···················· | ···················· | ···················· | NNNNNNNNNNNNNNNNNNNN | NNNNNNNNNNNNNNNNNNNN | 1 |
| I | 1654 | hCoV-19/USA/TX-HMH-6984/2020\|EPI_ISL_546408\|2020-0 | ···················· | ···················· | ···················· | ···················· | NNNNNNNNNNNNNNNNNNNN | NNNNNNNNNNNNNNNNNNNN | 1 |
| I | 1656 | hCoV-19/USA/TX-HMH-6986/2020\|EPI_ISL_546410\|2020-0 | ···················· | ···················· | ···················· | ···················· | NNNNNNNNNNNNNNNNNNNN | NNNNNNNNNNNNNNNNNNNN | 1 |
| I | 1657 | hCoV-19/USA/TX-HMH-6987/2020\|EPI_ISL_546411\|2020-0 | ···················· | ···················· | ···················· | ···················· | ········N····NNN···· | NNNN···NN·NN·NN·N··· | 1 |
| I | 1658 | hCoV-19/USA/TX-HMH-699/2020\|EPI_ISL_546413\|2020-04 | ···················· | ···················· | ···················· | ···················· | NNNNNNNNN··········· | ···················· | 1 |
| I | 1660 | hCoV-19/USA/TX-HMH-6997/2020\|EPI_ISL_546419\|2020-0 | ···················· | ···················· | ···················· | ···················· | NNNNNNNNNNNNNNNNNNNN | NNNNNNNNNNNNNNNNNNNN | 1 |
| I | 1661 | hCoV-19/USA/TX-HMH-7000/2020\|EPI_ISL_546421\|2020-0 | ···················· | ···················· | ···················· | ···················· | ·············N······ | NNNN··NN············ | 1 |
| I | 1663 | hCoV-19/USA/TX-HMH-7003/2020\|EPI_ISL_546424\|2020-0 | ···················· | ···················· | ···················· | ···················· | NNNNNNNNNNNNNNNNNNNN | NNNNNNNNNNNNNNNNNNNN | 1 |
| I | 1779 | hCoV-19/USA/CA-CZB-3327/2020\|EPI_ISL_548281\|2020-0 | ···················· | ·A·················· | ···················· | ···················· | NNNNNNNNNNNNNNNNNNNN | NNNNNNNNNNNNNNNNNNNN | 1 |
| I | 1780 | hCoV-19/USA/CA-CZB-3280/2020\|EPI_ISL_548282\|2020-0 | ···················· | ···················· | ···················· | ···················· | ······NNNNNNNNNNNNNN | NNNNNNNNNNNNNNNNNNNN | 1 |
| I | 1781 | hCoV-19/USA/CA-CZB-3348/2020\|EPI_ISL_548283\|2020-0 | ···················· | ···················· | ···················· | ···················· | ···················· | NNNNNNNNNNNNNNN····· | 1 |
| I | 1782 | hCoV-19/USA/CA-CZB-3296/2020\|EPI_ISL_548284\|2020-0 | ···················· | ···················· | ···················· | ···················· | ········NNNNNNNNNNNN | NNNNNNNNNNNNNNNNNNNN | 1 |
| I | 1784 | hCoV-19/USA/CA-CZB-3323/2020\|EPI_ISL_548286\|2020-0 | ···················· | ·A·················· | ···················· | ···················· | NNNNNNNNNNNNNNNNNNNN | NNNNNNNNNNNNNNNNNNNN | 1 |
| I | 1789 | hCoV-19/USA/CA-CZB-3324/2020\|EPI_ISL_548295\|2020-0 | ···················· | ·A·················· | ···················· | ···················· | ···················· | NNNNNNNNNNNNNNNNNNNN | 1 |
| I | 1796 | hCoV-19/USA/TX-HMH-5106/2020\|EPI_ISL_544529\|2020-0 | ···················· | ············A······· | ···················· | ···················· | NNNNNNNNNNNNNNNNNNNN | NNNNNNNNNNNNNNNNNNNN | 1 |
| I | 1797 | hCoV-19/USA/TX-HMH-5107/2020\|EPI_ISL_544530\|2020-0 | ···················· | ···················· | ···················· | ···················· | N····NNNN··N··N···NN | NN·N···NNN·········· | 1 |
| I | 1801 | hCoV-19/USA/TX-HMH-5117/2020\|EPI_ISL_544535\|2020-0 | ···················· | ············A······· | ···················· | ···················· | NNNNNNNNNNNNNNNNNNNN | NNNNNNNNNNNNNNNNNNNN | 1 |
| I | 1802 | hCoV-19/USA/TX-HMH-5118/2020\|EPI_ISL_544536\|2020-0 | ···················· | ···················· | ···················· | ···················· | ·············N······ | NN·N···N·······N···· | 1 |
| I | 1808 | hCoV-19/USA/TX-HMH-5130/2020\|EPI_ISL_544546\|2020-0 | ···················· | ···················· | ···················· | ···················· | NNNNNNNNNNNNNNNNNNNN | NNNNNNNNNNNNNNNNNNNN | 1 |
| I | 1810 | hCoV-19/USA/TX-HMH-5133/2020\|EPI_ISL_544549\|2020-0 | ···················· | ···················· | ···················· | ···················· | NNNNNNNNNNNNNNNNNNNN | NNNNNNNNNNNNNNNNNNNN | 1 |
| I | 1815 | hCoV-19/USA/TX-HMH-5150/2020\|EPI_ISL_544560\|2020-0 | ···················· | ···················· | ···················· | ···················· | NNNNNNNNNNNNNNNNNNNN | NNNNNNNNNNNNNNNNNNNN | 1 |
| I | 1818 | hCoV-19/USA/TX-HMH-5156/2020\|EPI_ISL_544565\|2020-0 | ···················· | NNNNNNNNNNNNNNNNNNNN | ···················· | ···················· | ···················· | ···················· | 1 |
| I | 1819 | hCoV-19/USA/TX-HMH-5157/2020\|EPI_ISL_544566\|2020-0 | ···················· | ···················· | ···················· | ···················· | ·············N······ | NNNN···NN··NN······· | 1 |
| I | 1820 | hCoV-19/USA/TX-HMH-5159/2020\|EPI_ISL_544567\|2020-0 | ···················· | ···················· | ···················· | ···················· | NNNNNNNNNNNNNNNNNNNN | NNNNNNNNNNNNNNNNNNNN | 1 |
| I | 1883 | hCoV-19/USA/TX-HMH-835/2020\|EPI_ISL_545858\|2020-04 | ···················· | ···················· | ···················· | ···················· | NNNNNNNNNNNNNNNNNNNN | NNNNNNNNNNNNNNNNNNNN | 1 |
| I | 1887 | hCoV-19/USA/TX-HMH-839/2020\|EPI_ISL_545862\|2020-04 | ···················· | ···················· | ···················· | ···················· | NNNNNNNNNNNNNNNNNNNN | NNNNNNNNNNNNNNNNNNNN | 1 |
| I | 1888 | hCoV-19/USA/TX-HMH-841/2020\|EPI_ISL_545863\|2020-04 | ···················· | ···················· | ···················· | ···················· | NNNNNNNNNNNNNNNNNNNN | NNNNNNNNNNNNNNNNNNNN | 1 |
| I | 1894 | hCoV-19/USA/TX-HMH-851/2020\|EPI_ISL_545871\|2020-04 | ···················· | ···················· | ···················· | ···················· | ·············N······ | NN·N···NN····NN····· | 1 |
| I | 1907 | hCoV-19/USA/TX-HMH-868/2020\|EPI_ISL_545885\|2020-04 | ···················· | ···················· | ···················· | ···················· | NNNNNNNNNNNNNNNNNNNN | NNNNNNNNNNNNNNNNNNNN | 1 |
| I | 1921 | hCoV-19/USA/TX-HMH-6873/2020\|EPI_ISL_546309\|2020-0 | ···················· | ···················· | ···················· | ···················· | NNNNNNNNNNNNNNNNNNNN | NNNNNNNNNNNNNNNNNNNN | 1 |
| I | 1928 | hCoV-19/USA/TX-HMH-6882/2020\|EPI_ISL_546317\|2020-0 | ···················· | ············A······· | ···················· | ···················· | NNNNNNNNNNNNNNNNNNNN | NNNNNNNNNNNNNNNNNNNN | 1 |
| I | 1929 | hCoV-19/USA/TX-HMH-6883/2020\|EPI_ISL_546318\|2020-0 | ···················· | ···················· | ···················· | ···················· | NNNNNNNNNNNNNNNNNNNN | NNNNNNNNNNNNNNNNNNNN | 1 |
| I | 1933 | hCoV-19/USA/TX-HMH-6888/2020\|EPI_ISL_546323\|2020-0 | ···················· | ···················· | ···················· | ···················· | NNNNNNNNNNNNNNNNNNNN | NNNNNNNNNNNNNNNNNNNN | 1 |
| I | 1936 | hCoV-19/USA/TX-HMH-6893/2020\|EPI_ISL_546327\|2020-0 | ···················· | ···················· | ···················· | ···················· | NNNNNNNNNNNNNNNNNNNN | NNNNNNNNNNNNNNNNNNNN | 1 |
| I | 1938 | hCoV-19/USA/TX-HMH-6895/2020\|EPI_ISL_546329\|2020-0 | ···················· | ···················· | ···················· | ···················· | NNNNNNNNNNNNNNNNNNNN | NNNNNNNNNNNNNNNNNNNN | 1 |
| I | 1939 | hCoV-19/USA/TX-HMH-6897/2020\|EPI_ISL_546331\|2020-0 | ···················· | ···················· | ···················· | ···················· | NNNNNNNNNNNNNNNNNNNN | NNNNNNNNNNNNNNNNNNNN | 1 |
| I | 1946 | hCoV-19/USA/TX-HMH-6903/2020\|EPI_ISL_546338\|2020-0 | ···················· | ···················· | ···················· | ···················· | NNNNNNNNNNNNNNNNNNNN | NNNNNNNNNNNNNNNNNNNN | 1 |
| I | 1947 | hCoV-19/USA/TX-HMH-6905/2020\|EPI_ISL_546339\|2020-0 | ···················· | ···················· | ···················· | ···················· | NNNNNNNNNNNNNNNNNNNN | NNNNNNNNNNNNNNNNNNNN | 1 |
| I | 1948 | hCoV-19/USA/TX-HMH-6907/2020\|EPI_ISL_546340\|2020-0 | ···················· | ·········A·········· | ···················· | ···················· | NNNNNNNNNNNNNNNNNNNN | NNNNNNNNNNNNNNNNNNNN | 1 |
| I | 1950 | hCoV-19/USA/TX-HMH-6910/2020\|EPI_ISL_546343\|2020-0 | ···················· | ···················· | ···················· | ···················· | NNNNNNNNNNNNNNNNNNNN | NNNNNNNNNNNNNNNNNNNN | 1 |
| I | 1951 | hCoV-19/USA/TX-HMH-6912/2020\|EPI_ISL_546345\|2020-0 | ···················· | ···················· | ···················· | ···················· | NNNNNNNNNNNNNNNNNNNN | NNNNNNNNNNNNNNNNNNNN | 1 |
| I | 1952 | hCoV-19/USA/TX-HMH-6915/2020\|EPI_ISL_546348\|2020-0 | ···················· | ···················· | ···················· | ···················· | NNNNNNNNNNNNNNNNNNNN | NNNNNNNNNNNNNNNNNNNN | 1 |
| I | 1954 | hCoV-19/USA/TX-HMH-6921/2020\|EPI_ISL_546353\|2020-0 | ···················· | ···················· | ···················· | ···················· | NNNNNNNNNNNNNNNNNNNN | NNNNNNNNNNNNNNNNNNNN | 1 |
| I | 1955 | hCoV-19/USA/TX-HMH-6923/2020\|EPI_ISL_546355\|2020-0 | ···················· | ···················· | ···················· | ···················· | NNNNNNNNNNNNNNNNNNNN | NNNNNNNNNNNNNNNNNNNN | 1 |
| I | 1956 | hCoV-19/USA/TX-HMH-6925/2020\|EPI_ISL_546356\|2020-0 | ···················· | ············A······· | ···················· | ···················· | NNNNNNNNNNNNNNNNNNNN | NNNNNNNNNNNNNNNNNNNN | 1 |
| I | 1957 | hCoV-19/USA/TX-HMH-6926/2020\|EPI_ISL_546357\|2020-0 | ···················· | ···················· | ···················· | ···················· | NNNNNNNNNNNNNNNNNNNN | NNNNNNNNNNNNNNNNNNNN | 1 |
| I | 1958 | hCoV-19/USA/TX-HMH-6930/2020\|EPI_ISL_546360\|2020-0 | ···················· | ···················· | ···················· | ···················· | NNNNNNNNNNNNNNNNNNNN | NNNNNNNNNNNNNNNNNNNN | 1 |
| I | 1960 | hCoV-19/USA/TX-HMH-6933/2020\|EPI_ISL_546362\|2020-0 | ···················· | ············A······· | ···················· | ···················· | NNNNNNNNNNNNNNNNNNNN | NNNNNNNNNNNNNNNNNNNN | 1 |
| I | 1963 | hCoV-19/USA/TX-HMH-6938/2020\|EPI_ISL_546366\|2020-0 | ···················· | ············A······· | ···················· | ···················· | NNNNNNNNNNNNNNNNNNNN | NNNNNNNNNNNNNNNNNNNN | 1 |
| I | 1965 | hCoV-19/USA/TX-HMH-6941/2020\|EPI_ISL_546370\|2020-0 | ···················· | ············A······· | ···················· | ···················· | NNNNNNNNNNNNNNNNNNNN | NNNNNNNNNNNNNNNNNNNN | 1 |
| I | 1968 | hCoV-19/USA/TX-HMH-6948/2020\|EPI_ISL_546376\|2020-0 | ···················· | ···················· | ···················· | ···················· | NNNNNNNNNNNNNNNNNNNN | NNNNNNNNNNNNNNNNNNNN | 1 |
| I | 1969 | hCoV-19/USA/TX-HMH-6950/2020\|EPI_ISL_546378\|2020-0 | ···················· | ···················· | ···················· | ···················· | NNNNNNNNNNNNNNNNNNNN | NNNNNNNNNNNNNNNNNNNN | 1 |
| I | 1971 | hCoV-19/USA/TX-HMH-6952/2020\|EPI_ISL_546380\|2020-0 | ···················· | ···················· | ···················· | ···················· | NNNNNNNN··NN··NN··NN | NN········NN········ | 1 |
| I | 2040 | hCoV-19/Canada/ON-PHL-20-01306/2020\|EPI_ISL_548909 | ···················· | ···················· | ···················· | ···················· | NNNNNNNNNNNNNNNNNNNN | NNNNNNNNNNNNNNN····· | 1 |
| I | 2043 | hCoV-19/Canada/ON-PHL-20-01309/2020\|EPI_ISL_548912 | ···················· | ···················· | ···················· | ···················· | ········NNNNNNNNNNNN | ···················· | 1 |
| I | 2052 | hCoV-19/USA/TX-HMH-5163/2020\|EPI_ISL_544570\|2020-0 | ···················· | ···················· | ···················· | ···················· | ·······N·····NN····· | NNNN······NN········ | 1 |
| I | 2053 | hCoV-19/USA/TX-HMH-5165/2020\|EPI_ISL_544571\|2020-0 | ···················· | ···················· | ···················· | ···················· | NNNNNNNNNNNNNNNNNNNN | NNNNNNNNNNNNNNNNNNNN | 1 |
| I | 2062 | hCoV-19/USA/TX-HMH-5182/2020\|EPI_ISL_544581\|2020-0 | ···················· | ···················· | ···················· | ···················· | NNNNNNNNNNNNNNNNNNNN | NNNNNNNNNNNNNNNNNNNN | 1 |
| I | 2063 | hCoV-19/USA/TX-HMH-5186/2020\|EPI_ISL_544583\|2020-0 | ···················· | ···················· | ···················· | ···················· | NNNNNNNNNNNNNNNNNNNN | NNNNNNNNNNNNNNNNNNNN | 1 |

```
I 2064 hCoV-19/USA/TX-HMH-5187/2020|EPI_ISL_544584|2020-0 ···················· ·············A······ ···················· ···················· NNNNNNNNNNNNNNNNNNNN NNNNNNNNNNNNNNNNNNNN 1
I 2066 hCoV-19/USA/TX-HMH-5191/2020|EPI_ISL_544586|2020-0 ···················· ···················· ···················· ···················· NNNNNNNNNNNNNNNNNNNN NNNNNNNNNNNNNNNNNNNN 1
I 2069 hCoV-19/USA/TX-HMH-5208/2020|EPI_ISL_544593|2020-0 ···················· ···················· ···················· ···················· ···N················ N······NN·······N··· 1
I 2070 hCoV-19/USA/TX-HMH-5209/2020|EPI_ISL_544594|2020-0 ···················· ···················· ···················· ···················· NNNNNNNNNNNNNNNNNNNN NNNNNNNNNNNNNNNNNNNN 1
I 2073 hCoV-19/USA/TX-HMH-5211/2020|EPI_ISL_544597|2020-0 ···················· ···················· ···················· ···················· ···N·······N········ NNNN···NNNNN·NN····N 1
I 2074 hCoV-19/USA/TX-HMH-5213/2020|EPI_ISL_544598|2020-0 ···················· ·············A······ ···················· ···················· ········N··········· NN·N···NN·N········· 1
I 2077 hCoV-19/USA/TX-HMH-5219/2020|EPI_ISL_544603|2020-0 ···················· ···················· ···················· ···················· NNNNNNNNNNNNNNNNNNNN NNNNNNNNNNNNNNNNNNNN 1
I 2079 hCoV-19/USA/TX-HMH-5220/2020|EPI_ISL_544605|2020-0 ···················· ···················· ···················· ···················· NNNNNNNNNNNNNNNNNNNN NNNNNNNNNNNNNNNNNNNN 1
I 2081 hCoV-19/USA/TX-HMH-5230/2020|EPI_ISL_544609|2020-0 ···················· ···················· ···················· ···················· NNNNNNNNNNNNNNNNNNNN NNNNNNNNNNNNNNNNNNNN 1
I 2083 hCoV-19/USA/TX-HMH-5233/2020|EPI_ISL_544612|2020-0 ···················· ···················· ···················· ···················· NNNNNNNNNNNNNNNNNNNN NNNNNNNNNNNNNNNNNNNN 1
I 2085 hCoV-19/USA/TX-HMH-5236/2020|EPI_ISL_544614|2020-0 ···················· ···················· ···················· ···················· NNNNNNNNNNNNNNNNNNNN NNNNNNNNNNNNNNNNNNNN 1
I 2086 hCoV-19/USA/TX-HMH-5237/2020|EPI_ISL_544615|2020-0 ···················· ···················· ···················· ···················· NNNNNNNNNNNNNNNNNNNN NNNNNNNNNNNNNNNNNNNN 1
I 2087 hCoV-19/USA/TX-HMH-524/2020|EPI_ISL_544617|2020-04 ···················· ···················· ···················· ···················· ···N·NNNN·····NN···· N··N··NNNNNNNNN····· 1
I 2088 hCoV-19/USA/TX-HMH-5242/2020|EPI_ISL_544619|2020-0 ···················· ···················· ···················· ···················· NNNNNNNNNNNNNNNNNNNN NNNNNNNNNNNNNNNNNNNN 1
I 2090 hCoV-19/USA/TX-HMH-5245/2020|EPI_ISL_544622|2020-0 ···················· ···················· ···················· ···················· NNNNNNNNNNNNNNNNNNNN NNNNNNNNNNNNNNNNNNNN 1
I 2092 hCoV-19/USA/TX-HMH-5247/2020|EPI_ISL_544624|2020-0 ···················· ······A············· ···················· ···················· NNNNN···NN·········· NNNN···NN····N····N· 1
I 2093 hCoV-19/USA/TX-HMH-5250/2020|EPI_ISL_544627|2020-0 ···················· ···················· ···················· ···················· NNNNNNNNNNNNNNNNNNNN NNNNNNNNNNNNNNNNNNNN 1
I 2094 hCoV-19/USA/TX-HMH-5254/2020|EPI_ISL_544629|2020-0 ···················· ·············A······ ···················· ···················· NNNNNNNNNNNNNNNNNNNN NNNNNNNNNNNNNNNNNNNN 1
I 2096 hCoV-19/USA/TX-HMH-5260/2020|EPI_ISL_544633|2020-0 ···················· ···················· ···················· ···················· NNNNNNNNNNNNNNNNNNNN NNNNNNNNNNNNNNNNNNNN 1
I 2097 hCoV-19/USA/TX-HMH-5261/2020|EPI_ISL_544634|2020-0 ···················· ···················· ···················· ···················· NNNNNNNNNNNNNNNNNNNN NNNNNNNNNNNNNNNNNNNN 1
I 2100 hCoV-19/USA/TX-HMH-5269/2020|EPI_ISL_544640|2020-0 ···················· ···················· ···················· ···················· NNNNNNNNNNNNNNNNNNNN NNNNNNNNNNNNNNNNNNNN 1
I 2103 hCoV-19/USA/TX-HMH-5273/2020|EPI_ISL_544643|2020-0 ···················· ···················· ···················· ···················· ···N·····NNN··NNNNN· NN·N···NN·NN·NN·N·N· 1
I 2148 hCoV-19/USA/TX-HMH-913/2020|EPI_ISL_545914|2020-04 ···················· ···················· ···················· ···················· NNNNNNNNNNNNNNN····· ···················· 1
I 2151 hCoV-19/USA/TX-HMH-916/2020|EPI_ISL_545917|2020-04 ···················· ···················· ···················· ···················· NNNNNNNNNNNNNNNNNNNN NNNNNNNNNNNNNNNNNNNN 1
I 2156 hCoV-19/USA/TX-HMH-928/2020|EPI_ISL_545923|2020-04 ···················· ···················· ···················· ···················· NNNNNNNNNNNNNNNNNNNN NNNNNNNNNNNNNNNNNNNN 1
I 2162 hCoV-19/USA/TX-HMH-956/2020|EPI_ISL_545930|2020-04 ···················· ···················· ···················· ···················· NNNNNNNNNNNNNNNNNNNN ···················· 1
I 2175 hCoV-19/USA/TX-HMH-978/2020|EPI_ISL_545943|2020-04 ···················· ···················· ···················· ···················· NNNNNNNNNNNNNNNNNNNN NNNNNNNNNNNNNNNNNNNN 1
I 2187 hCoV-19/USA/TX-HMH-6384/2020|EPI_ISL_545958|2020-0 ···················· ···················· ···················· ···················· NNNNNNNNNNNNNNNNNNNN NNNNNNNNNNNNNNNNNNNN 1
I 2195 hCoV-19/USA/TX-HMH-6397/2020|EPI_ISL_545970|2020-0 ···················· ···················· ···················· ···················· NNNNNNNNNNNNNNNNNNNN NNNNNNNNNNNNNNNNNNNN 1
I 2197 hCoV-19/USA/TX-HMH-6403/2020|EPI_ISL_545974|2020-0 ···················· ···················· ···················· ···················· ··············N····· NN·N···NN····N······ 1
I 2199 hCoV-19/USA/TX-HMH-6866/2020|EPI_ISL_546301|2020-0 ···················· ···················· ···················· ···················· ···················· NN·N···NN··········· 1
I 2201 hCoV-19/USA/TX-HMH-6868/2020|EPI_ISL_546303|2020-0 ···················· ···················· ···················· ···················· NNNNN··NN··N··NNNN·· NNNN·········NNN···· 1
I 2204 hCoV-19/USA/TX-HMH-6964/2020|EPI_ISL_546391|2020-0 ···················· ·············A······ ···················· ···················· NNNNNNNNNNNNNNNNNNNN NNNNNNNNNNNNNNNNNNNN 1
I 2206 hCoV-19/USA/TX-HMH-6970/2020|EPI_ISL_546396|2020-0 ···················· ···················· ···················· ···················· NNNNNNNNNNNNNNNNNNNN NNNNNNNNNNNNNNNNNNNN 1
I 2208 hCoV-19/USA/TX-HMH-6974/2020|EPI_ISL_546399|2020-0 ···················· ···················· ···················· ···················· NNNNNNNNNNNNNNNNNNNN NNNNNNNNNNNNNNNNNNNN 1
I 2249 hCoV-19/New Zealand/20CV0152/2020|EPI_ISL_548108|2 ···················· ···················· ···················· ···················· NNNNNNNNNNNNNNNNNNNN NNNNNNNNNNNNNNNNNNNN 1
I 2282 hCoV-19/Canada/ON-PHL-20-01310/2020|EPI_ISL_548913 ···················· ···················· ···················· ···················· NNNNNNNNNNNNNNNNNNNN NNNNNNNNNNNNNNNNNNNN 1
I 2284 hCoV-19/Canada/ON-PHL-20-01313/2020|EPI_ISL_548916 ···················· ···················· ···················· ···················· ···················· NNNNNNNNNNNNNNNN···· 1
I 2301 hCoV-19/Canada/ON-PHL-20-90089/2020|EPI_ISL_548938 ···················· ···················· ···················· ···················· ···················· NNNNNNNNNNNNNN······ 1
I 2312 hCoV-19/USA/TX-HMH-4935/2020|EPI_ISL_544411|2020-0 ···················· ···················· ···················· ···················· NNNNNNNNNNNNNNNNNNNN NNNNNNNNNNNNNNNNNNNN 1
I 2314 hCoV-19/USA/TX-HMH-4939/2020|EPI_ISL_544414|2020-0 ···················· ···················· ···················· ···················· NNNNNNNNNNNNNNNNNNNN NNNNNNNNNNNNNNNNNNNN 1
I 2323 hCoV-19/USA/TX-HMH-5056/2020|EPI_ISL_544492|2020-0 ···················· NNNNNNNNNNNNNNNNNNNN ···················· ···················· ···················· ···················· 1
I 2332 hCoV-19/USA/TX-HMH-7952/2020|EPI_ISL_545735|2020-0 ···················· ···················· ···················· ···················· NNNNNNNNNNNNNNNNNNNN NNNNNNNNNNNNNNNNNNNN 1
I 2333 hCoV-19/USA/TX-HMH-7953/2020|EPI_ISL_545736|2020-0 ···················· ···················· ···················· ···················· NNNNNNNNNNNNNNNNNNNN NNNNNNNNNNNNNNNNNNNN 1
I 2334 hCoV-19/USA/TX-HMH-7955/2020|EPI_ISL_545737|2020-0 ···················· ···················· ···················· ···················· ··············N····· ·······NN····NN····· 1
I 2335 hCoV-19/USA/TX-HMH-7956/2020|EPI_ISL_545738|2020-0 ···················· ···················· ···················· ···················· NNNNNNNNNNNNNNNNNNNN NNNNNNNNNNNNNNNNNNNN 1
I 2337 hCoV-19/USA/TX-HMH-7958/2020|EPI_ISL_545740|2020-0 ···················· ···················· ···················· ···················· NNNNNNNNNNNNNNNNNNNN NNNNNNNNNNNNNNNNNNNN 1
I 2340 hCoV-19/USA/TX-HMH-7964/2020|EPI_ISL_545746|2020-0 ···················· ·············A······ ···················· ···················· NNNNNNNNNNNNNNNNNNNN NNNNNNNNNNNNNNNNNNNN 1
I 2342 hCoV-19/USA/TX-HMH-7967/2020|EPI_ISL_545749|2020-0 ···················· ·A·················· ···················· ···················· NNNNNNNNNNNNNNNNNNNN NNNNNNNNNNNNNNNNNNNN 1
I 2345 hCoV-19/USA/TX-HMH-7971/2020|EPI_ISL_545753|2020-0 ···················· ·A·················· ···················· ···················· NNNNNNNNNNNNNNNNNNNN NNNNNNNNNNNNNNNNNNNN 1
I 2346 hCoV-19/USA/TX-HMH-7972/2020|EPI_ISL_545754|2020-0 ···················· ·A·················· ···················· ···················· NNNNNNNNNNNNNNNNNNNN NNNNNNNNNNNNNNNNNNNN 1
I 2347 hCoV-19/USA/TX-HMH-7973/2020|EPI_ISL_545755|2020-0 ···················· ···················· ···················· ···················· NNNNNNNNNNNNNNNNNNNN NNNNNNNNNNNNNNNNNNNN 1
I 2352 hCoV-19/USA/TX-HMH-6739/2020|EPI_ISL_546206|2020-0 ···················· ···················· ···················· ···················· NN·N······N···NN·N·· NNNN···NN·NN·NNN···· 1
I 2356 hCoV-19/USA/TX-HMH-6745/2020|EPI_ISL_546210|2020-0 ···················· ···················· ···················· ···················· ···················· NN·····NN·····N····· 1
I 2358 hCoV-19/USA/TX-HMH-6747/2020|EPI_ISL_546212|2020-0 ···················· ···················· ···················· ···················· NNNNNNNNNNNNNNNNNNNN NNNNNNNNNNNNNNNNNNNN 1
I 2360 hCoV-19/USA/TX-HMH-675/2020|EPI_ISL_546214|2020-04 ···················· ···················· ···················· ···················· NNNNNNNNNNNNNNNNNNNN ···················· 1
I 2368 hCoV-19/USA/TX-HMH-6761/2020|EPI_ISL_546224|2020-0 ···················· ···················· ···················· ···················· NNNNNNNNNNNNNNNNNNNN NNNNNNNNNNNNNNNNNNNN 1
I 2370 hCoV-19/USA/TX-HMH-6763/2020|EPI_ISL_546226|2020-0 ···················· ···················· ···················· ···················· NNNNNNNNNNNNNNNNNNNN NNNNNNNNNNNNNNNNNNNN 1
I 2372 hCoV-19/USA/TX-HMH-6765/2020|EPI_ISL_546228|2020-0 ···················· ···················· ···················· ···················· NNNNNNNNNNNNNNNNNNNN NNNNNNNNNNNNNNNNNNNN 1
I 2380 hCoV-19/USA/TX-HMH-6781/2020|EPI_ISL_546239|2020-0 ···················· ···················· ···················· ···················· NNNNNNNNNNNNNNNNNNNN NNNNNNNNNNNNNNNNNNNN 1
I 2387 hCoV-19/USA/TX-HMH-6792/2020|EPI_ISL_546246|2020-0 ···················· ···················· ···················· ···················· NNNNNNNNNNNNNNNNNNNN NNNNNNNNNNNNNNNNNNNN 1
I 2390 hCoV-19/USA/TX-HMH-6798/2020|EPI_ISL_546250|2020-0 ···················· ···················· ···················· ···················· NNNNNNNNNNNNNNNNNNNN NNNNNNNNNNNNNNNNNNNN 1
I 2400 hCoV-19/USA/TX-HMH-6810/2020|EPI_ISL_546260|2020-0 ···················· ···················· ···················· ···················· NNNNNNNNNNNNNNNNNNNN NNNNNNNNNNNNNNNNNNNN 1
I 2402 hCoV-19/USA/TX-HMH-6816/2020|EPI_ISL_546263|2020-0 ···················· ···················· ···················· ···················· ·NNN··········N····· NNNN······NN······N· 1
I 2404 hCoV-19/USA/TX-HMH-6820/2020|EPI_ISL_546265|2020-0 ···················· ···················· ···················· ···················· NNNNNNNNNNNNNNNNNNNN NNNNNNNNNNNNNNNNNNNN 1
I 2405 hCoV-19/USA/TX-HMH-6822/2020|EPI_ISL_546267|2020-0 ···················· ···················· ···················· ···················· NNNNNNNNNNNNNNNNNNNN NNNNNNNNNNNNNNNNNNNN 1
I 2406 hCoV-19/USA/TX-HMH-6824/2020|EPI_ISL_546269|2020-0 ···················· ···················· ···················· ···················· NNNNNNNNNNNNNNNNNNNN NNNNNNNNNNNNNNNNNNNN 1
I 2500 hCoV-19/Switzerland/GE-SNRCI-2432/2020|EPI_ISL_548 ···················· ···················A ···················· ···················· ·······NNNNNNNNNNNNN ···················· 1
I 2510 hCoV-19/Switzerland/GE-SNRCI-3080/2020|EPI_ISL_548 ···················· ·A·················· ···················· ···················· ·······NNNNNNNNNNNNN ···················· 1
I 2519 hCoV-19/Canada/ON-PHL-20-01229/2020|EPI_ISL_548851 ···················· ···················· ···················· ···················· NNNNNNNNNNNNNNNNNNNN ···················· 1
I 2524 hCoV-19/Canada/ON-PHL-20-01234/2020|EPI_ISL_548856 ···················· ···················· ···················· ···················· NNNN················ ···················· 1
I 2533 hCoV-19/Canada/ON-PHL-20-01244/2020|EPI_ISL_548866 ···················· ···················· ···················· ···················· NNNNNNNNNNNNNNNNNNNN NNNNNNNNNNNNNNNNNNNN 1
I 2538 hCoV-19/Canada/ON-PHL-20-01250/2020|EPI_ISL_548872 ···················· ···················· ···················· ···················· NNNNNNNNNNNNNNNNNNNN NNNNNNNNNNNNNNNNNNNN 1
I 2539 hCoV-19/Canada/ON-PHL-20-01255/2020|EPI_ISL_548873 ···················· ···················· ···················· ···················· ···················· NNNNNNNNNNNNNNNNNNNN 1
I 2541 hCoV-19/Canada/ON-PHL-20-01260/2020|EPI_ISL_548875 ···················· ···················· ···················· ···················· NNNNNNN············· ···················· 1
I 2543 hCoV-19/Canada/ON-PHL-20-01263/2020|EPI_ISL_548877 ···················· ···················· ···················· ···················· NNNNNNNNNNNNNNNNNNNN NNNNNNNNNNNNNNNNNNNN 1
I 2546 hCoV-19/Canada/ON-PHL-20-01273/2020|EPI_ISL_548881 ···················· ···················· ···················· ···················· NNNNN········N······ ···················· 1
I 2555 hCoV-19/Canada/ON-PHL-20-01288/2020|EPI_ISL_548893 ···················· ···················· ···················· ···················· NNNNNNNNNNNNNNNN···· ···················· 1
I 2559 hCoV-19/Canada/ON-PHL-20-01294/2020|EPI_ISL_548898 ···················· ···················· ···················· ···················· NNNNNNNNNNNNNNNNNNNN NNNNNNNNNNNNNNNNNNNN 1
I 2562 hCoV-19/USA/TX-HMH-4952/2020|EPI_ISL_544426|2020-0 ···················· ···················· ···················· ···················· NNNNNNNNNNNNNNNNNNNN NNNNNNNNNNNNNNNNNNNN 1
```

| | | | | | | | | |
|---|---|---|---|---|---|---|---|---|
| I | 2566 hCoV-19/USA/TX-HMH-4958/2020\|EPI_ISL_544432\|2020-0 | ···················· | ···················· | ···················· | ···················· | NNNNNNNNNNNNNNNNNNNN | NNNNNNNNNNNNNNNNNNNN | 1 |
| I | 2573 hCoV-19/USA/TX-HMH-4990/2020\|EPI_ISL_544441\|2020-0 | ···················· | ···················· | ···················· | ···················· | NNNNNNNNNNNNNNNNNNNN | NNNNNNNNNNNNNNNNNNNN | 1 |
| I | 2574 hCoV-19/USA/TX-HMH-4993/2020\|EPI_ISL_544443\|2020-0 | ···················· | ···················· | ···················· | ···················· | NNNNNNNNNNNNNNNNNNNN | NNNNNNNNNNNNNNNNNNNN | 1 |
| I | 2575 hCoV-19/USA/TX-HMH-4996/2020\|EPI_ISL_544445\|2020-0 | ···················· | ···················· | ···················· | ···················· | NNNNNNNNNNNNNNNNNNNN | NNNNNNNNNNNNNNNNNNNN | 1 |
| I | 2576 hCoV-19/USA/TX-HMH-4998/2020\|EPI_ISL_544446\|2020-0 | ···················· | ···················· | ···················· | ···················· | NNNNNNNNNNNNNNNNNNNN | NNNNNNNNNNNNNNNNNNNN | 1 |
| I | 2577 hCoV-19/USA/TX-HMH-4999/2020\|EPI_ISL_544447\|2020-0 | ···················· | ···················· | ···················· | ···················· | NNNNNNNNNNNNNNNNNNNN | NNNNNNNNNNNNNNNNNNNN | 1 |
| I | 2578 hCoV-19/USA/TX-HMH-5005/2020\|EPI_ISL_544452\|2020-0 | ···················· | ···················· | ···················· | ···················· | NNNNNNNNNNNNNNNNNNNN | NNNNNNNNNNNNNNNNNNNN | 1 |
| I | 2582 hCoV-19/USA/TX-HMH-5012/2020\|EPI_ISL_544458\|2020-0 | ···················· | ···················· | ···················· | ···················· | NNNNNNNNNNNNNNNNNNNN | NNNNNNNNNNNNNNNNNNNN | 1 |
| I | 2583 hCoV-19/USA/TX-HMH-5013/2020\|EPI_ISL_544459\|2020-0 | ···················· | ···················· | ···················· | ···················· | NNNNNNNNNNNNNNNNNNNN | NNNNNNNNNNNNNNNNNNNN | 1 |
| I | 2589 hCoV-19/USA/TX-HMH-5021/2020\|EPI_ISL_544467\|2020-0 | ···················· | ···················· | ···················· | ···················· | NNNNNNNNNNNNNNNNNNNN | NNNNNNNNNNNNNNNNNNNN | 1 |
| I | 2590 hCoV-19/USA/TX-HMH-5025/2020\|EPI_ISL_544468\|2020-0 | ···················· | ···················· | ···················· | ···················· | ···················· | NN·····NN··········· | 1 |
| I | 2596 hCoV-19/USA/TX-HMH-5035/2020\|EPI_ISL_544475\|2020-0 | ···················· | NNNNNNNNNNNNNNNNNNNN | ···················· | ···················· | ···················· | ···················· | 1 |
| I | 2604 hCoV-19/USA/TX-HMH-5049/2020\|EPI_ISL_544484\|2020-0 | ···················· | ·············A······ | ···················· | ···················· | NNNNNNNNNNNNNNNNNNNN | NNNNNNNNNNNNNNNNNNNN | 1 |
| I | 2618 hCoV-19/USA/TX-HMH-5087/2020\|EPI_ISL_544515\|2020-0 | ···················· | ···················· | ···················· | ···················· | NNNNNNNNNNNNNNNNNNNN | NNNNNNNNNNNNNNNNNNNN | 1 |
| I | 2623 hCoV-19/USA/TX-HMH-5095/2020\|EPI_ISL_544521\|2020-0 | ···················· | ···················· | ···················· | ···················· | NNNNNNNNNNNNNNNNNNNN | NNNNNNNNNNNNNNNNNNNN | 1 |
| I | 2626 hCoV-19/USA/TX-HMH-7974/2020\|EPI_ISL_545756\|2020-0 | ···················· | ···················· | ···················· | ···················· | NNNNNNNNNNNNNNNNNNNN | NNNNNNNNNNNNNNNNNNNN | 1 |
| I | 2631 hCoV-19/USA/TX-HMH-7981/2020\|EPI_ISL_545762\|2020-0 | ···················· | ·A·················· | ···················· | ···················· | ··············NN···· | NNN··········N····NN | 1 |
| I | 2632 hCoV-19/USA/TX-HMH-7982/2020\|EPI_ISL_545763\|2020-0 | ···················· | ···················· | ···················· | ···················· | NNNNNNNNNNNNNNNNNNNN | NNNNNNNNNNNNNNNNNNNN | 1 |
| I | 2633 hCoV-19/USA/TX-HMH-7983/2020\|EPI_ISL_545764\|2020-0 | ···················· | ···················· | ···················· | ···················· | ···N··········N···NN | NNNN···N·N·····N··N· | 1 |
| I | 2644 hCoV-19/USA/TX-HMH-7999/2020\|EPI_ISL_545779\|2020-0 | ···················· | ··············C····· | ···················· | ···················· | NNNNNNNNNNNNNNNNNNNN | NNNNNNNNNNNNNNNNNNNN | 1 |
| I | 2658 hCoV-19/USA/TX-HMH-8018/2020\|EPI_ISL_545795\|2020-0 | ···················· | ···················· | ···················· | ···················· | NNNNNNNNNNNNNNNNNNNN | NNNNNNNNNNNNNNNNNNNN | 1 |
| I | 2662 hCoV-19/USA/TX-HMH-8027/2020\|EPI_ISL_545799\|2020-0 | ···················· | ···················· | ···················· | ···················· | NNNNNNNNNNNNNNNNNNNN | NNNNNNNNNNNNNNNNNNNN | 1 |
| I | 2670 hCoV-19/USA/TX-HMH-8038/2020\|EPI_ISL_545808\|2020-0 | ···················· | ·············A······ | ···················· | ···················· | NNNNNNNNNNNNNNNNNNNN | NNNNNNNNNNNNNNNNNNNN | 1 |
| I | 2675 hCoV-19/USA/TX-HMH-8043/2020\|EPI_ISL_545814\|2020-0 | ···················· | ·············A······ | ···················· | ···················· | NNNNNNNNNNNNNNNNNNNN | NNNNNNNNNNNNNNNNNNNN | 1 |
| I | 2676 hCoV-19/USA/TX-HMH-8046/2020\|EPI_ISL_545815\|2020-0 | ···················· | ···················· | ···················· | ···················· | NNNNNNNNNNNNNNNNNNNN | NNNNNNNNNNNNNNNNNNNN | 1 |
| I | 2686 hCoV-19/USA/TX-HMH-8060/2020\|EPI_ISL_545827\|2020-0 | ···················· | ···················· | ···················· | ···················· | NNNNNNNNNNNNNNNNNNNN | NNNNNNNNNNNNNNNNNNNN | 1 |
| I | 2688 hCoV-19/USA/TX-HMH-8063/2020\|EPI_ISL_545829\|2020-0 | ···················· | ···················· | ···················· | ···················· | NNNNNNNNNNNNNNNNNNNN | NNNNNNNNNNNNNNNNNNNN | 1 |
| I | 2695 hCoV-19/USA/TX-HMH-8070/2020\|EPI_ISL_545836\|2020-0 | ···················· | ···················· | ···················· | ···················· | NNNNNNNNNNNNNNNNNNNN | NNNNNNNNNNNNNNNNNNNN | 1 |
| I | 2712 hCoV-19/USA/TX-HMH-6837/2020\|EPI_ISL_546277\|2020-0 | ···················· | ···················· | ···················· | ···················· | NNNNNNNNNNNNNNNNNNNN | NNNNNNNNNNNNNNNNNNNN | 1 |
| I | 2713 hCoV-19/USA/TX-HMH-6838/2020\|EPI_ISL_546278\|2020-0 | ···················· | ·············A······ | ···················· | ···················· | NNNNNNNNNNNNNNNNNNNN | NNNNNNNNNNNNNNNNNNNN | 1 |
| I | 2714 hCoV-19/USA/TX-HMH-6839/2020\|EPI_ISL_546279\|2020-0 | ···················· | ···················· | ···················· | ···················· | NNNNNNNNNNNNNNNNNNNN | NNNNNNNNNNNNNNNNNNNN | 1 |
| I | 2716 hCoV-19/USA/TX-HMH-6843/2020\|EPI_ISL_546282\|2020-0 | ···················· | ···················· | ···················· | ···················· | NNNNNNNNNNNNNNNNNNNN | NNNNNNNNNNNNNNNNNNNN | 1 |
| I | 2717 hCoV-19/USA/TX-HMH-6845/2020\|EPI_ISL_546284\|2020-0 | ···················· | ···················· | ···················· | ···················· | NNNNNNNNNNNNNNNNNNNN | NNNNNNNNNNNNNNNNNNNN | 1 |
| I | 2718 hCoV-19/USA/TX-HMH-6849/2020\|EPI_ISL_546287\|2020-0 | ···················· | ···················· | ···················· | ···················· | NNNNNNNNNNNNNNNNNNNN | NNNNNNNNNNNNNNNNNNNN | 1 |
| I | 2721 hCoV-19/USA/TX-HMH-6853/2020\|EPI_ISL_546291\|2020-0 | ···················· | ·············A······ | ···················· | ···················· | NNNNNNNNNNNNNNNNNNNN | NNNNNNNNNNNNNNNNNNNN | 1 |
| I | 2722 hCoV-19/USA/TX-HMH-6854/2020\|EPI_ISL_546292\|2020-0 | ···················· | ···················· | ···················· | ···················· | NNNNNNNNNNNNNNNNNNNN | NNNNNNNNNNNNNNNNNNNN | 1 |
| I | 2723 hCoV-19/USA/TX-HMH-6855/2020\|EPI_ISL_546293\|2020-0 | ···················· | ···················· | ···················· | ···················· | NNNNNNNNNNNNNNNNNNNN | NNNNNNNNNNNNNNNNNNNN | 1 |
| I | 2817 hCoV-19/USA/TX-HMH-4743/2020\|EPI_ISL_544300\|2020-0 | ···················· | ···················· | ···················· | ···················· | NNNNNNNNNNNNNNNNNNNN | NNNNNNNNNNNNNNNNNNNN | 1 |
| I | 2818 hCoV-19/USA/TX-HMH-4748/2020\|EPI_ISL_544302\|2020-0 | ···················· | ···················· | ···················· | ···················· | NNNNNNNNNNNNNNNNNNNN | NNNNNNNNNNNNNNNNNNNN | 1 |
| I | 2819 hCoV-19/USA/TX-HMH-4749/2020\|EPI_ISL_544303\|2020-0 | ···················· | ··················NN | ···················· | ···················· | NNNNNNNNNNNNNNNNNNNN | NNNNNNNNNNNNNNNNNNNN | 1 |
| I | 2820 hCoV-19/USA/TX-HMH-4750/2020\|EPI_ISL_544304\|2020-0 | ···················· | ···················· | ···················· | ···················· | NNNNNNNNNNNNNNNNNNNN | NNNNNNNNNNNNNNNNNNNN | 1 |
| I | 2821 hCoV-19/USA/TX-HMH-4752/2020\|EPI_ISL_544306\|2020-0 | ···················· | ···················· | ···················· | ···················· | NNNNNNNNNNNNNNNNNNNN | NNNNNNNNNNNNNNNNNNNN | 1 |
| I | 2822 hCoV-19/USA/TX-HMH-4753/2020\|EPI_ISL_544307\|2020-0 | ···················· | ···················· | ···················· | ···················· | NNNNNNNNNNNNNNNNNNNN | NNNNNNNNNNNNNNNNNNNN | 1 |
| I | 2823 hCoV-19/USA/TX-HMH-4754/2020\|EPI_ISL_544308\|2020-0 | ···················· | ···················· | ···················· | ···················· | NNNNNNNNNNNNNNNNNNNN | NNNNNNNNNNNNNNNNNNNN | 1 |
| I | 2825 hCoV-19/USA/TX-HMH-4759/2020\|EPI_ISL_544310\|2020-0 | ···················· | ···················· | ···················· | ···················· | NNNNNNNNNNNNNNNNNNNN | NNNNNNNNNNNNNNNNNNNN | 1 |
| I | 2827 hCoV-19/USA/TX-HMH-4760/2020\|EPI_ISL_544312\|2020-0 | ···················· | ···················· | ···················· | ···················· | NNNNNNNNNNNNNNNNNNNN | NNNNNNNNNNNNNNNNNNNN | 1 |
| I | 2845 hCoV-19/USA/TX-HMH-4872/2020\|EPI_ISL_544393\|2020-0 | ···················· | NNNNNNNNNNNNNNNNNNNN | ···················· | ···················· | ···················· | ···················· | 1 |
| I | 2850 hCoV-19/USA/TX-HMH-4880/2020\|EPI_ISL_544399\|2020-0 | ···················· | ···················· | ···················· | ···················· | NNNNNNNNNNNNNNNNNNNN | NNNNNNNNNNNNNNNNNNNN | 1 |
| I | 2855 hCoV-19/USA/TX-HMH-7766/2020\|EPI_ISL_545624\|2020-0 | ···················· | ···················· | ···················· | ···················· | NNNNNNNNNNNNNNNNNNNN | NNNNNNNNNNNNNNNNNNNN | 1 |
| I | 2890 hCoV-19/USA/TX-HMH-6637/2020\|EPI_ISL_546135\|2020-0 | ···················· | ·············A······ | ···················· | ···················· | NNNNNNNNNNNNNNNNNNNN | NNNNNNNNNNNNNNNNNNNN | 1 |
| I | 2895 hCoV-19/USA/TX-HMH-6643/2020\|EPI_ISL_546140\|2020-0 | ···················· | ···················· | ···················· | ···················· | NNNNNNNNNNNNNNNNNNNN | NNNNNNNNNNNNNNNNNNNN | 1 |
| I | 2896 hCoV-19/USA/TX-HMH-6645/2020\|EPI_ISL_546141\|2020-0 | ···················· | ···················· | ···················· | ···················· | NNNNNNNNNNNNNNNNNNNN | NNNNNNNNNNNNNNNNNNNN | 1 |
| I | 2897 hCoV-19/USA/TX-HMH-6648/2020\|EPI_ISL_546144\|2020-0 | ···················· | ···················· | ···················· | ···················· | NNNNNNNNNNNNNNNNNNNN | NNNNNNNNNNNNNNNNNNNN | 1 |
| I | 2898 hCoV-19/USA/TX-HMH-6649/2020\|EPI_ISL_546145\|2020-0 | ···················· | ···················· | ···················· | ···················· | NNNNNNNNNNNNNNNNNNNN | NNNNNNNNNNNNNNNNNNNN | 1 |
| I | 2902 hCoV-19/USA/TX-HMH-6654/2020\|EPI_ISL_546149\|2020-0 | ···················· | ·············A······ | ···················· | ···················· | NNNNNNNNNNNNNNNNNNNN | NNNNNNNNNNNNNNNNNNNN | 1 |
| I | 2903 hCoV-19/USA/TX-HMH-5685/2020\|EPI_ISL_546945\|2020-0 | ···················· | ···················· | ···················· | ···················· | NNNNNNNNNNNNNNNNNNNN | NNNNNNNNNNNNNNNNNNNN | 1 |
| I | 2904 hCoV-19/USA/TX-HMH-5689/2020\|EPI_ISL_546946\|2020-0 | ···················· | ···················· | ···················· | ···················· | NNNNNNNNNNNNNNNNNNNN | NNNNNNNNNNNNNNNNNNNN | 1 |
| I | 2905 hCoV-19/USA/TX-HMH-5692/2020\|EPI_ISL_546948\|2020-0 | ···················· | ·············A······ | ···················· | ···················· | NNNNNNNNNNNNNNNNNNNN | NNNNNNNNNNNNNNNNNNNN | 1 |
| I | 2908 hCoV-19/USA/TX-HMH-5698/2020\|EPI_ISL_546951\|2020-0 | ···················· | ······A············· | ···················· | ···················· | NNNNNNNNNNNNNNNNNNNN | NNNNNNNNNNNNNNNNNNNN | 1 |
| I | 2910 hCoV-19/USA/TX-HMH-5701/2020\|EPI_ISL_546953\|2020-0 | ···················· | ···················· | ···················· | ···················· | NNNNNNNNNNNNNNNNNNNN | NNNNNNNNNNNNNNNNNNNN | 1 |
| I | 2911 hCoV-19/USA/TX-HMH-5702/2020\|EPI_ISL_546954\|2020-0 | ···················· | ···················· | ···················· | ···················· | NNNNNNNNNNNNNNNNNNNN | NNNNNNNNNNNNNNNNNNNN | 1 |
| I | 2915 hCoV-19/USA/TX-HMH-5709/2020\|EPI_ISL_546958\|2020-0 | ···················· | ···················· | ···················· | ···················· | NNNNNNNNNNNNNNNNNNNN | NNNNNNNNNNNNNNNNNNNN | 1 |
| I | 2916 hCoV-19/USA/TX-HMH-5711/2020\|EPI_ISL_546961\|2020-0 | ···················· | ···················· | ···················· | ···················· | NNNNNNNNNNNNNNNNNNNN | NNNNNNNNNNNNNNNNNNNN | 1 |
| I | 2917 hCoV-19/USA/TX-HMH-5712/2020\|EPI_ISL_546962\|2020-0 | ···················· | ···················· | ···················· | ···················· | NNNNNNNNNNNNNNNNNNNN | NNNNNNNNNNNNNNNNNNNN | 1 |
| I | 2918 hCoV-19/USA/TX-HMH-5717/2020\|EPI_ISL_546963\|2020-0 | ···················· | ···················· | ···················· | ···················· | NNNNNNNNNNNNNNNNNNNN | NNNNNNNNNNNNNNNNNNNN | 1 |
| I | 2919 hCoV-19/USA/TX-HMH-5718/2020\|EPI_ISL_546964\|2020-0 | ···················· | ·············A······ | ···················· | ···················· | NNNNNNNNNNNNNNNNNNNN | NNNNNNNNNNNNNNNNNNNN | 1 |
| I | 2920 hCoV-19/USA/TX-HMH-5720/2020\|EPI_ISL_546966\|2020-0 | ···················· | ···················· | ···················· | ···················· | NNNNNNNNNNNNNNNNNNNN | NNNNNNNNNNNNNNNNNNNN | 1 |
| I | 2925 hCoV-19/USA/TX-HMH-5734/2020\|EPI_ISL_546975\|2020-0 | ···················· | ···················· | ···················· | ···················· | NNNNNNNNNNNNNNNNNNNN | NNNNNNNNNNNNNNNNNNNN | 1 |
| I | 2926 hCoV-19/USA/TX-HMH-5735/2020\|EPI_ISL_546976\|2020-0 | ···················· | ···················· | ···················· | ···················· | NNNNNNNNNNNNNNNNNNNN | NNNNNNNNNNNNNNNNNNNN | 1 |
| I | 2935 hCoV-19/USA/TX-HMH-6355/2020\|EPI_ISL_547412\|2020-0 | ···················· | ···················· | ···················· | ···················· | NNNNNNNNNNNNNNNNNNNN | NNNNNNNNNNNNNNNNNNNN | 1 |
| I | 2937 hCoV-19/USA/TX-HMH-6357/2020\|EPI_ISL_547414\|2020-0 | ···················· | ···················· | ···················· | ···················· | NNNNNNNNNNNNNNNNNNNN | NNNNNNNNNNNNNNNNNNNN | 1 |
| I | 2939 hCoV-19/USA/TX-HMH-6361/2020\|EPI_ISL_547418\|2020-0 | ···················· | ·············A······ | ···················· | ···················· | NNNNNNNNNNNNNNNNNNNN | NNNNNNNNNNNNNNNNNNNN | 1 |
| I | 2940 hCoV-19/USA/TX-HMH-6362/2020\|EPI_ISL_547419\|2020-0 | ···················· | ···················· | ···················· | ···················· | NNNNNNNNNNNNNNNNNNNN | NNNNNNNNNNNNNNNNNNNN | 1 |
| I | 3059 hCoV-19/Canada/ON-PHL-20-01163/2020\|EPI_ISL_548785 | ···················· | ···················· | ···················· | ···················· | ·NNNNNNNNNNNNNNNNNNN | ···················· | 1 |
| I | 3078 hCoV-19/USA/TX-HMH-2981/2020\|EPI_ISL_543909\|2020-0 | ···················· | ···················· | ···················· | ···················· | NNNNNNNNNNNNNNNNNNNN | NNNNNNNNNNNNNNNNNNNN | 1 |
| I | 3079 hCoV-19/USA/TX-HMH-2982/2020\|EPI_ISL_543910\|2020-0 | ···················· | ···················· | ···················· | ···················· | NNNNNNNNNNNNNNNNNNNN | NNNNNNNNNNNNNNNNNNNN | 1 |
| I | 3082 hCoV-19/USA/TX-HMH-2992/2020\|EPI_ISL_543916\|2020-0 | ···················· | ··············C····· | ···················· | ···················· | NNNNNNNNNNNNNNNNNNNN | NNNNNNNNNNNNNNNNNNNN | 1 |
| I | 3090 hCoV-19/USA/TX-HMH-3050/2020\|EPI_ISL_543956\|2020-0 | ···················· | ···················· | ···················· | ···················· | NNNNNNNNNNNNNNNNNNNN | NNNNNNNNNNNNNNNNNNNN | 1 |
| I | 3092 hCoV-19/USA/TX-HMH-3053/2020\|EPI_ISL_543959\|2020-0 | ···················· | ···················· | ···················· | ···················· | NNNNNNNNNNNNNNNNNNNN | NNNNNNNNNNNNNNNNNNNN | 1 |
| I | 3096 hCoV-19/USA/TX-HMH-3060/2020\|EPI_ISL_543966\|2020-0 | ···················· | ······A············· | ···················· | ···················· | NNNNNNNNNNNNNNNNNNNN | NNNNNNNNNNNNNNNNNNNN | 1 |

I 3105 hCoV-19/USA/TX-HMH-3080/2020|EPI_ISL_543981|2020-0 ···················· ···················· ···················· ···················· ··N·····N·····NN···· NN·N···NN·NN·NN····· 1
I 3116 hCoV-19/USA/TX-HMH-3108/2020|EPI_ISL_543997|2020-0 ···················· ···················· ···················· ···················· ········NN···NN····· NN·N···NN··········· 1
I 3119 hCoV-19/USA/TX-HMH-4765/2020|EPI_ISL_544315|2020-0 ···················· ···················· ···················· ···················· NNNNNNNNNNNNNNNNNNNN NNNNNNNNNNNNNNNNNNNN 1
I 3120 hCoV-19/USA/TX-HMH-4766/2020|EPI_ISL_544316|2020-0 ···················· ·············A······ ···················· ···················· NNNNNNNNNNNNNNNNNNNN NNNNNNNNNNNNNNNNNNNN 1
I 3122 hCoV-19/USA/TX-HMH-4769/2020|EPI_ISL_544319|2020-0 ···················· ···················· ···················· ···················· NNNNNNNNNNNNNNNNNNNN NNNNNNNNNNNNNNNNNNNN 1
I 3123 hCoV-19/USA/TX-HMH-4773/2020|EPI_ISL_544320|2020-0 ···················· ···················· ···················· ···················· NNNNNNNNNNNNNNNNNNNN NNNNNNNNNNNNNNNNNNNN 1
I 3124 hCoV-19/USA/TX-HMH-4774/2020|EPI_ISL_544321|2020-0 ···················· ···················· ···················· ···················· NNNNNNNNNNNNNNNNNNNN NNNNNNNNNNNNNNNNNNNN 1
I 3125 hCoV-19/USA/TX-HMH-4777/2020|EPI_ISL_544323|2020-0 ···················· ···················· ···················· ···················· NNNNNNNNNNNNNNNNNNNN NNNNNNNNNNNNNNNNNNNN 1
I 3126 hCoV-19/USA/TX-HMH-4779/2020|EPI_ISL_544324|2020-0 ···················· ···················· ···················· ···················· NNNNNNNNNNNNNNNNNNNN NNNNNNNNNNNNNNNNNNNN 1
I 3128 hCoV-19/USA/TX-HMH-4784/2020|EPI_ISL_544326|2020-0 ···················· ·············A······ ···················· ···················· NNNNNNNNNNNNNNNNNNNN NNNNNNNNNNNNNNNNNNNN 1
I 3129 hCoV-19/USA/TX-HMH-4787/2020|EPI_ISL_544329|2020-0 ···················· ···················· ···················· ···················· NNNNNNNNNNNNNNNNNNNN NNNNNNNNNNNNNNNNNNNN 1
I 3130 hCoV-19/USA/TX-HMH-4788/2020|EPI_ISL_544330|2020-0 ···················· ···················· ···················· ···················· NNNNNNNNNNNNNNNNNNNN NNNNNNNNNNNNNNNNNNNN 1
I 3131 hCoV-19/USA/TX-HMH-4789/2020|EPI_ISL_544331|2020-0 ···················· ···················· ···················· ···················· NNNNNNNNNNNNNNNNNNNN NNNNNNNNNNNNNNNNNNNN 1
I 3133 hCoV-19/USA/TX-HMH-4790/2020|EPI_ISL_544333|2020-0 ···················· ···················· ···················· ···················· NNNNNNNNNNNNNNNNNNNN NNNNNNNNNNNNNNNNNNNN 1
I 3135 hCoV-19/USA/TX-HMH-4792/2020|EPI_ISL_544335|2020-0 ···················· ···················· ···················· ···················· NNNNNNNNNNNNNNNNNNNN NNNNNNNNNNNNNNNNNNNN 1
I 3136 hCoV-19/USA/TX-HMH-4798/2020|EPI_ISL_544337|2020-0 ···················· ···················· ···················· ···················· NNNNNNNNNNNNNNNNNNNN NNNNNNNNNNNNNNNNNNNN 1
I 3138 hCoV-19/USA/TX-HMH-4804/2020|EPI_ISL_544342|2020-0 ···················· ···················· ···················· ···················· NNNNNNNNNNNNNNNNNNNN NNNNNNNNNNNNNNNNNNNN 1
I 3139 hCoV-19/USA/TX-HMH-4808/2020|EPI_ISL_544344|2020-0 ···················· ···················· ···················· ···················· NNNNNNNNNNNNNNNNNNNN NNNNNNNNNNNNNNNNNNNN 1
I 3141 hCoV-19/USA/TX-HMH-4810/2020|EPI_ISL_544346|2020-0 ···················· ···················· ···················· ···················· NNNNNNNNNNNNNNNNNNNN NNNNNNNNNNNNNNNNNNNN 1
I 3142 hCoV-19/USA/TX-HMH-4814/2020|EPI_ISL_544348|2020-0 ···················· ···················· ···················· ···················· NNNNNNNNNNNNNNNNNNNN NNNNNNNNNNNNNNNNNNNN 1
I 3143 hCoV-19/USA/TX-HMH-4815/2020|EPI_ISL_544349|2020-0 ···················· ···················· ···················· ···················· ············NNNN·N·· NN··N··NN·N··NN····· 1
I 3155 hCoV-19/USA/TX-HMH-4833/2020|EPI_ISL_544363|2020-0 ···················· ···················· ···················· ···················· ··············N····· NN·····NN··········· 1
I 3162 hCoV-19/USA/TX-HMH-4881/2020|EPI_ISL_544400|2020-0 ···················· ···················· ···················· ···················· NNNNNNNNNNNNNNNNNNNN NNNNNNNNNNNNNNNNNNNN 1
I 3163 hCoV-19/USA/TX-HMH-4883/2020|EPI_ISL_544402|2020-0 ···················· NNNNNNNNNNNNNNNNNNNN ···················· ···················· ···················· ···················· 1
I 3185 hCoV-19/USA/TX-HMH-7871/2020|EPI_ISL_545674|2020-0 ···················· ···················· ···················· ···················· NNNNNNNNNNNNNNNNNNNN NNNNNNNNNNNNNNNNNNNN 1
I 3194 hCoV-19/USA/TX-HMH-7887/2020|EPI_ISL_545683|2020-0 ···················· ·············A······ ···················· ···················· ··············N····· NN·N···NN··········· 1
I 3197 hCoV-19/USA/TX-HMH-7894/2020|EPI_ISL_545686|2020-0 ···················· ···················· ···················· ···················· NNNNNNNNNNNNNNNNNNNN NNNNNNNNNNNNNNNNNNNN 1
I 3199 hCoV-19/USA/TX-HMH-7896/2020|EPI_ISL_545688|2020-0 ···················· ···················· ···················· ···················· NNNNNNNNNNNNNNNNNNNN NNNNNNNNNNNNNNNNNNNN 1
I 3203 hCoV-19/USA/TX-HMH-7901/2020|EPI_ISL_545693|2020-0 ···················· ···················· ···················· ···················· NNNNNNNNNNNNNNNNNNNN NNNNNNNNNNNNNNNNNNNN 1
I 3204 hCoV-19/USA/TX-HMH-7903/2020|EPI_ISL_545694|2020-0 ···················· ···················· ···················· ···················· NNNNNNNNNNNNNNNNNNNN NNNNNNNNNNNNNNNNNNNN 1
I 3206 hCoV-19/USA/TX-HMH-7906/2020|EPI_ISL_545696|2020-0 ···················· ······A············· ···················· ···················· NNNNNNNNNNNNNNNNNNNN NNNNNNNNNNNNNNNNNNNN 1
I 3210 hCoV-19/USA/TX-HMH-7912/2020|EPI_ISL_545701|2020-0 ···················· ···················· ···················· ···················· NNNNNNNNNNNNNNNNNNNN NNNNNNNNNNNNNNNNNNNN 1
I 3211 hCoV-19/USA/TX-HMH-7913/2020|EPI_ISL_545702|2020-0 ···················· ···················· ···················· ···················· NNNNNNNNNNNNNNNNNNNN NNNNNNNNNNNNNNNNNNNN 1
I 3212 hCoV-19/USA/TX-HMH-7915/2020|EPI_ISL_545703|2020-0 ···················· ···················· ···················· ···················· NNNNNNNNNNNNNNNNNNNN NNNNNNNNNNNNNNNNNNNN 1
I 3213 hCoV-19/USA/TX-HMH-7916/2020|EPI_ISL_545704|2020-0 ···················· ···················· ···················· ···················· NNNNNNNNNNNNNNNNNNNN NNNNNNNNNNNNNNNNNNNN 1
I 3214 hCoV-19/USA/TX-HMH-7918/2020|EPI_ISL_545706|2020-0 ···················· ···················· ···················· ···················· NNNNNNNNNNNNNNNNNNNN NNNNNNNNNNNNNNNNNNNN 1
I 3217 hCoV-19/USA/TX-HMH-7923/2020|EPI_ISL_545709|2020-0 ···················· ···················· ···················· ···················· NNNNNNNNNNNNNNNNNNNN NNNNNNNNNNNNNNNNNNNN 1
I 3218 hCoV-19/USA/TX-HMH-7924/2020|EPI_ISL_545710|2020-0 ···················· ···················· ···················· ···················· NNNNNNNNNNNNNNNNNNNN NNNNNNNNNNNNNNNNNNNN 1
I 3219 hCoV-19/USA/TX-HMH-7927/2020|EPI_ISL_545712|2020-0 ···················· ·············A······ ···················· ···················· NNNNNNNNNNNNNNNNNNNN NNNNNNNNNNNNNNNNNNNN 1
I 3221 hCoV-19/USA/TX-HMH-7930/2020|EPI_ISL_545715|2020-0 ···················· ···················· ···················· ···················· NNNNNNNNNNNNNNNNNNNN NNNNNNNNNNNNNNNNNNNN 1
I 3222 hCoV-19/USA/TX-HMH-7931/2020|EPI_ISL_545716|2020-0 ···················· ···················· ···················· ···················· ··············N···NN NN·N···NN··N········ 1
I 3223 hCoV-19/USA/TX-HMH-7932/2020|EPI_ISL_545717|2020-0 ···················· ···················· ···················· ···················· NNNNNNNNNNNNNNNNNNNN NNNNNNNNNNNNNNNNNNNN 1
I 3224 hCoV-19/USA/TX-HMH-7933/2020|EPI_ISL_545718|2020-0 ···················· ···················· ···················· ···················· NNNNNNNNNNNNNNNNNNNN NNNNNNNNNNNNNNNNNNNN 1
I 3231 hCoV-19/USA/TX-HMH-7940/2020|EPI_ISL_545725|2020-0 ···················· N·················N· ···················· ···················· NNNNNNNNNNNNNNNNNNNN NNNNNNNNNNNNNNNNNNNN 1
I 3232 hCoV-19/USA/TX-HMH-7944/2020|EPI_ISL_545727|2020-0 ···················· ···················· ···················· ···················· NNNNNNNNNNNNNNNNNNNN NNNNNNNNNNNNNNNNNNNN 1
I 3234 hCoV-19/USA/TX-HMH-7947/2020|EPI_ISL_545730|2020-0 ···················· ···················· ···················· ···················· ·············NNN··NN NN···········NN····· 1
I 3235 hCoV-19/USA/TX-HMH-7949/2020|EPI_ISL_545731|2020-0 ···················· ···················· ···················· ···················· NNNNNNNNNNNNNNNNNNNN NNNNNNNNNNNNNNNNNNNN 1
I 3237 hCoV-19/USA/TX-HMH-7950/2020|EPI_ISL_545733|2020-0 ···················· ···················· ···················· ···················· NNNNNNNNNNNNNNNNNNNN NNNNNNNNNNNNNNNNNNNN 1
I 3238 hCoV-19/USA/TX-HMH-6655/2020|EPI_ISL_546150|2020-0 ···················· ···················· ···················· ···················· NNNNNNNNNNNNNNNNNNNN NNNNNNNNNNNNNNNNNNNN 1
I 3239 hCoV-19/USA/TX-HMH-6657/2020|EPI_ISL_546151|2020-0 ···················· ···················· ···················· ···················· NNNNNNNNNNNNNNNNNNNN NNNNNNNNNNNNNNNNNNNN 1
I 3240 hCoV-19/USA/TX-HMH-6661/2020|EPI_ISL_546154|2020-0 ···················· ···················· ···················· ···················· NNNNNNNNNNNNNNNNNNNN NNNNNNNNNNNNNNNNNNNN 1
I 3241 hCoV-19/USA/TX-HMH-6662/2020|EPI_ISL_546155|2020-0 ···················· ···················· ···················· ···················· NNNNNNNNNNNNNNNNNNNN NNNNNNNNNNNNNNNNNNNN 1
I 3242 hCoV-19/USA/TX-HMH-6664/2020|EPI_ISL_546156|2020-0 ···················· ···················· ···················· ···················· NNNNNNNNNNNNNNNNNNNN NNNNNNNNNNNNNNNNNNNN 1
I 3244 hCoV-19/USA/TX-HMH-6667/2020|EPI_ISL_546158|2020-0 ···················· ···················· ···················· ···················· NNNNNNNNNNNNNNNNNNNN NNNNNNNNNNNNNNNNNNNN 1
I 3245 hCoV-19/USA/TX-HMH-6670/2020|EPI_ISL_546160|2020-0 ···················· ···················· ···················· ···················· NNNNNNNNNNNNNNNNNNNN NNNNNNNNNNNNNNNNNNNN 1
I 3246 hCoV-19/USA/TX-HMH-6673/2020|EPI_ISL_546162|2020-0 ···················· ···················· ···················· ···················· NNNNNNNNNNNNNNNNNNNN NNNNNNNNNNNNNNNNNNNN 1
I 3247 hCoV-19/USA/TX-HMH-6675/2020|EPI_ISL_546164|2020-0 ···················· ···················· ···················· ···················· NNNNNNNNNNNNNNNNNNNN NNNNNNNNNNNNNNNNNNNN 1
I 3248 hCoV-19/USA/TX-HMH-6677/2020|EPI_ISL_546165|2020-0 ···················· ···················· ···················· ···················· NNNNNNNNNNNNNNNNNNNN NNNNNNNNNNNNNNNNNNNN 1
I 3249 hCoV-19/USA/TX-HMH-6680/2020|EPI_ISL_546168|2020-0 ···················· ···················· ···················· ···················· NNNNNNNNNNNNNNNNNNNN NNNNNNNNNNNNNNNNNNNN 1
I 3250 hCoV-19/USA/TX-HMH-6682/2020|EPI_ISL_546170|2020-0 ···················· ···················· ···················· ···················· NNNNNNNNNNNNNNNNNNNN NNNNNNNNNNNNNNNNNNNN 1
I 3251 hCoV-19/USA/TX-HMH-6683/2020|EPI_ISL_546171|2020-0 ···················· ···················· ···················· ···················· NNNNNNNNNNNNNNNNNNNN NNNNNNNNNNNNNNNNNNNN 1
I 3252 hCoV-19/USA/TX-HMH-6685/2020|EPI_ISL_546172|2020-0 ···················· ···················· ···················· ···················· NNNNNNNNNNNNNNNNNNNN NNNNNNNNNNNNNNNNNNNN 1
I 3253 hCoV-19/USA/TX-HMH-6686/2020|EPI_ISL_546173|2020-0 ···················· ···················· ···················· ···················· NNNNNNNNNNNNNNNNNNNN NNNNNNNNNNNNNNNNNNNN 1
I 3254 hCoV-19/USA/TX-HMH-6697/2020|EPI_ISL_546177|2020-0 ···················· ·············A······ ···················· ···················· NNNNNNNNNNNNNNNNNNNN NNNNNNNNNNNNNNNNNNNN 1
I 3257 hCoV-19/USA/TX-HMH-6701/2020|EPI_ISL_546180|2020-0 ···················· ···················· ···················· ···················· NNNNNNNNNNNNNNNNNNNN NNNNNNNNNNNNNNNNNNNN 1
I 3261 hCoV-19/USA/TX-HMH-6708/2020|EPI_ISL_546185|2020-0 ···················· ···················· ···················· ···················· NNNNNNNNNNNNNNNNNNNN NNNNNNNNNNNNNNNNNNNN 1
I 3264 hCoV-19/USA/TX-HMH-6718/2020|EPI_ISL_546191|2020-0 ···················· ···················· ···················· ···················· NNNNNNNNNNNNNNNNNNNN NNNNNNNNNNNNNNNNNNNN 1
I 3265 hCoV-19/USA/TX-HMH-6720/2020|EPI_ISL_546192|2020-0 ···················· ···················· ···················· ···················· NNNNNNNNNNNNNNNNNNNN NNNNNNNNNNNNNNNNNNNN 1
I 3268 hCoV-19/USA/TX-HMH-6725/2020|EPI_ISL_546195|2020-0 ···················· ···················· ···················· ···················· NNNNNNNNNNNNNNNNNNNN NNNNNNNNNNNNNNNNNNNN 1
I 3271 hCoV-19/USA/TX-HMH-673/2020|EPI_ISL_546198|2020-04 ···················· ···················· ···················· ···················· NNNNNNNNNNNNNNNNNNNN ···················· 1
I 3272 hCoV-19/USA/TX-HMH-6730/2020|EPI_ISL_546199|2020-0 ···················· ···················· ···················· ···················· NNNNNNNNNNNNNNNNNNNN NNNNNNNNNNNNNNNNNNNN 1
I 3273 hCoV-19/USA/TX-HMH-5736/2020|EPI_ISL_546977|2020-0 ···················· ···················· ···················· ···················· NNNNNNNNNNNNNNNNNNNN NNNNNNNNNNNNNNNNNNNN 1
I 3276 hCoV-19/USA/TX-HMH-5741/2020|EPI_ISL_546981|2020-0 ···················· ·············A······ ···················· ···················· NNNNNNNNNNNNNNNNNNNN NNNNNNNNNNNNNNNNNNNN 1
I 3279 hCoV-19/USA/TX-HMH-5747/2020|EPI_ISL_546985|2020-0 ···················· ···················· ···················· ···················· NNNNNNNNNNNNNNNNNNNN NNNNNNNNNNNNNNNNNNNN 1
I 3286 hCoV-19/USA/TX-HMH-5762/2020|EPI_ISL_546996|2020-0 ···················· ···················· ···················· ···················· NNNNNNNNNNNNNNNNNNNN NNNNNNNNNNNNNNNNNNNN 1
I 3329 hCoV-19/USA/TX-HMH-2823/2020|EPI_ISL_543798|2020-0 ···················· ···················· ···················· ···················· NNNNNNNNNNNNNNNNNNNN NNNNNNNNNNNNNNNNNNNN 1
I 3332 hCoV-19/USA/TX-HMH-3004/2020|EPI_ISL_543926|2020-0 ···················· ···················· ···················· ···················· NNNNNNNNNNNNNNNNNNNN NNNNNNNNNNNNNNNNNNNN 1
I 3333 hCoV-19/USA/TX-HMH-3006/2020|EPI_ISL_543927|2020-0 ···················· ···················· ···················· ···················· NNNNNNNNNNNNNNNNNNNN NNNNNNNNNNNNNNNNNNNN 1
I 3341 hCoV-19/USA/TX-HMH-3021/2020|EPI_ISL_543939|2020-0 ···················· ···················· ···················· ···················· ···N····NNN···N····· NN·N·NNNNNNNNNN····· 1
I 3353 hCoV-19/USA/TX-HMH-4688/2020|EPI_ISL_544257|2020-0 ···················· ···················· ···················· ···················· NNNNNNNNNNNNNNNNNNNN NNNNNNNNNNNNNNNNNNNN 1

| | | | | | | | | |
|---|---|---|---|---|---|---|---|---|
| I | 3355 hCoV-19/USA/TX-HMH-4691/2020\|EPI_ISL_544261\|2020-0 | ···················· | ···················· | ···················· | ···················· | NNNNNNNNNNNNNNNNNNNN | NNNNNNNNNNNNNNNNNNNN | 1 |
| I | 3358 hCoV-19/USA/TX-HMH-4699/2020\|EPI_ISL_544267\|2020-0 | ···················· | ···················· | ···················· | ···················· | NNNNNNNNNNNNNNNNNNNN | NNNNNNNNNNNNNNNNNNNN | 1 |
| I | 3360 hCoV-19/USA/TX-HMH-4702/2020\|EPI_ISL_544270\|2020-0 | ···················· | ···················· | ···················· | ···················· | NNNNNNNNNNNNNNNNNNNN | NNNNNNNNNNNNNNNNNNNN | 1 |
| I | 3362 hCoV-19/USA/TX-HMH-4705/2020\|EPI_ISL_544273\|2020-0 | ···················· | ···················· | ···················· | ···················· | NNNNNNNNNNNNNNNNNNNN | NNNNNNNNNNNNNNNNNNNN | 1 |
| I | 3364 hCoV-19/USA/TX-HMH-4709/2020\|EPI_ISL_544277\|2020-0 | ···················· | ···················· | ···················· | ···················· | NNNNNNNNNNNNNNNNNNNN | NNNNNNNNNNNNNNNNNNNN | 1 |
| I | 3370 hCoV-19/USA/TX-HMH-4719/2020\|EPI_ISL_544284\|2020-0 | ···················· | ···················· | ···················· | ···················· | NNNNNNNNNNNNNNNNNNNN | NNNNNNNNNNNNNNNNNNNN | 1 |
| I | 3372 hCoV-19/USA/TX-HMH-4720/2020\|EPI_ISL_544286\|2020-0 | ···················· | ···················· | ···················· | ···················· | NNNNNNNNNNNNNNNNNNNN | NNNNNNNNNNNNNNNNNNNN | 1 |
| I | 3373 hCoV-19/USA/TX-HMH-4725/2020\|EPI_ISL_544288\|2020-0 | ···················· | ···················· | ···················· | ···················· | NNNNNNNNNNNNNNNNNNNN | NNNNNNNNNNNNNNNNNNNN | 1 |
| I | 3374 hCoV-19/USA/TX-HMH-4729/2020\|EPI_ISL_544290\|2020-0 | ···················· | ···················· | ···················· | ···················· | NNNNNNNNNNNNNNNNNNNN | NNNNNNNNNNNNNNNNNNNN | 1 |
| I | 3375 hCoV-19/USA/TX-HMH-4735/2020\|EPI_ISL_544294\|2020-0 | ···················· | ···················· | ···················· | ···················· | NNNNNNNNNNNNNNNNNNNN | NNNNNNNNNNNNNNNNNNNN | 1 |
| I | 3377 hCoV-19/USA/TX-HMH-4740/2020\|EPI_ISL_544297\|2020-0 | ···················· | ···················· | ···················· | ···················· | NNNNNNNNNNNNNNNNNNNN | NNNNNNNNNNNNNNNNNNNN | 1 |
| I | 3378 hCoV-19/USA/TX-HMH-4741/2020\|EPI_ISL_544298\|2020-0 | ···················· | ···················· | ···················· | ···················· | NNNNNNNNNNNNNNNNNNNN | NNNNNNNNNNNNNNNNNNNN | 1 |
| I | 3379 hCoV-19/USA/TX-HMH-4742/2020\|EPI_ISL_544299\|2020-0 | ···················· | ···················· | ···················· | ···················· | NNNNNNNNNNNNNNNNNNNN | NNNNNNNNNNNNNNNNNNNN | 1 |
| I | 3380 hCoV-19/USA/TX-HMH-2717/2020\|EPI_ISL_545500\|2020-0 | ···················· | ···················· | ···················· | ···················· | NNNNNNNNNNNNNNNNNNNN | NNNNNNNNNNNNNNNNNNNN | 1 |
| I | 3401 hCoV-19/USA/TX-HMH-2747/2020\|EPI_ISL_545524\|2020-0 | ···················· | ···················· | ···················· | ···················· | NNNNNNNNNNNNNNNNNNNN | NNNNNNNNNNNNNNNNNNNN | 1 |
| I | 3406 hCoV-19/USA/TX-HMH-7685/2020\|EPI_ISL_545584\|2020-0 | ···················· | ···················· | ···················· | ···················· | NNNNNNNNNNNNNNNNNNNN | NNNNNNNNNNNNNNNNNNNN | 1 |
| I | 3407 hCoV-19/USA/TX-HMH-7688/2020\|EPI_ISL_545586\|2020-0 | ···················· | ·············A······ | ···················· | ···················· | NNNNNNNNNNNNNNNNNNNN | NNNNNNNNNNNNNNNNNNNN | 1 |
| I | 3409 hCoV-19/USA/TX-HMH-7695/2020\|EPI_ISL_545588\|2020-0 | ···················· | ···················· | ···················· | ···················· | NNNNNNNNNNNNNNNNNNNN | NNNNNNNNNNNNNNNNNNNN | 1 |
| I | 3415 hCoV-19/USA/TX-HMH-7711/2020\|EPI_ISL_545597\|2020-0 | ···················· | ···················· | ···················· | ···················· | NNNNNNNNNNNNNNNNNNNN | NNNNNNNNNNNNNNNNNNNN | 1 |
| I | 3418 hCoV-19/USA/TX-HMH-6446/2020\|EPI_ISL_546001\|2020-0 | ···················· | ···················· | ···················· | ···················· | NNNNNNNNNNNNNNNNNNNN | NNNNNNNNNNNNNNNNNNNN | 1 |
| I | 3419 hCoV-19/USA/TX-HMH-6447/2020\|EPI_ISL_546002\|2020-0 | ···················· | ·············A······ | ···················· | ···················· | NNNNNNNNNNNNNNNNNNNN | NNNNNNNNNNNNNNNNNNNN | 1 |
| I | 3420 hCoV-19/USA/TX-HMH-6450/2020\|EPI_ISL_546004\|2020-0 | ···················· | ·············A······ | ···················· | ···················· | NNNNNNNNNNNNNNNNNNNN | NNNNNNNNNNNNNNNNNNNN | 1 |
| I | 3428 hCoV-19/USA/TX-HMH-6460/2020\|EPI_ISL_546013\|2020-0 | ···················· | ···················· | ···················· | ···················· | NNNNNNNNNNNNNNNNNNNN | NNNNNNNNNNNNNNNNNNNN | 1 |
| I | 3431 hCoV-19/USA/TX-HMH-6468/2020\|EPI_ISL_546018\|2020-0 | ···················· | ···················· | ···················· | ···················· | NNNNNNNNNNNNNNNNNNNN | NNNNNNNNNNNNNNNNNNNN | 1 |
| I | 3436 hCoV-19/USA/TX-HMH-6478/2020\|EPI_ISL_546024\|2020-0 | ···················· | ···················· | ···················· | ···················· | NNNNNNNNNNNNNNNNNNNN | NNNNNNNNNNNNNNNNNNNN | 1 |
| I | 3444 hCoV-19/USA/TX-HMH-7531/2020\|EPI_ISL_546826\|2020-0 | ···················· | ···················· | ···················· | ···················· | NNNNNNNNNNNNNNNNNNNN | NNNNNNNNNNNNNNNNNNNN | 1 |
| I | 3451 hCoV-19/USA/TX-HMH-7543/2020\|EPI_ISL_546838\|2020-0 | ···················· | ···················· | ···················· | ···················· | ··N·NNNN·····NNNNN·· | NN·····NN··········· | 1 |
| I | 3452 hCoV-19/USA/TX-HMH-7544/2020\|EPI_ISL_546839\|2020-0 | ···················· | ···················· | ···················· | ···················· | NNNNNNNNNNNNNNNNNNNN | NNNNNNNNNNNNNNNNNNNN | 1 |
| I | 3453 hCoV-19/USA/TX-HMH-7545/2020\|EPI_ISL_546840\|2020-0 | ···················· | ···················· | ···················· | ···················· | ·NNNNN··NN····NNNN·· | NNNN···NNN···N·NN··· | 1 |
| I | 3457 hCoV-19/USA/TX-HMH-7551/2020\|EPI_ISL_546844\|2020-0 | ···················· | ·············A······ | ···················· | ···················· | ···················· | NN·····NN··········· | 1 |
| I | 3458 hCoV-19/USA/TX-HMH-7552/2020\|EPI_ISL_546845\|2020-0 | ···················· | ···················· | ···················· | ···················· | NNNNNNNNNNNNNNNNNNNN | NNNNNNNNNNNNNNNNNNNN | 1 |
| I | 3459 hCoV-19/USA/TX-HMH-7555/2020\|EPI_ISL_546847\|2020-0 | ···················· | ···················· | ···················· | ···················· | NNNNNNNNNNNNNNNNNNNN | NNNNNNNNNNNNNNNNNNNN | 1 |
| I | 3461 hCoV-19/USA/TX-HMH-7560/2020\|EPI_ISL_546852\|2020-0 | ···················· | ···················· | ···················· | ···················· | NNNNNNNNNNNNNNNNNNNN | NNNNNNNNNNNNNNNNNNNN | 1 |
| I | 3462 hCoV-19/USA/TX-HMH-7561/2020\|EPI_ISL_546853\|2020-0 | ···················· | ···················· | ···················· | ···················· | ··············N····· | NN·····NN··········· | 1 |
| I | 3463 hCoV-19/USA/TX-HMH-7562/2020\|EPI_ISL_546854\|2020-0 | ···················· | ···················· | ···················· | ···················· | NNNNNNNNNNNNNNNNNNNN | NNNNNNNNNNNNNNNNNNNN | 1 |
| I | 3464 hCoV-19/USA/TX-HMH-7564/2020\|EPI_ISL_546855\|2020-0 | ···················· | ···················· | ···················· | ···················· | NNNNNNNNNNNNNNNNNNNN | NNNNNNNNNNNNNNNNNNNN | 1 |
| I | 3469 hCoV-19/USA/TX-HMH-6193/2020\|EPI_ISL_547303\|2020-0 | ···················· | ·············A······ | ···················· | ···················· | NNNNNNNNNNNNNNNNNNNN | NNNNNNNNNNNNNNNNNNNN | 1 |
| I | 3470 hCoV-19/USA/TX-HMH-6195/2020\|EPI_ISL_547304\|2020-0 | ···················· | ···················· | ···················· | ···················· | NNNNNNNNNNNNNNNNNNNN | NNNNNNNNNNNNNNNNNNNN | 1 |
| I | 3472 hCoV-19/USA/TX-HMH-6202/2020\|EPI_ISL_547309\|2020-0 | ···················· | ···················· | ···················· | ···················· | NNNNNNNNNNNNNNNNNNNN | NNNNNNNNNNNNNNNNNNNN | 1 |
| I | 3481 hCoV-19/USA/TX-HMH-6217/2020\|EPI_ISL_547320\|2020-0 | ···················· | ···················· | ···················· | ···················· | NNNNNNNNNNNNNNNNNNNN | NNNNNNNNNNNNNNNNNNNN | 1 |
| I | 3483 hCoV-19/USA/TX-HMH-6219/2020\|EPI_ISL_547322\|2020-0 | ···················· | ···················· | ···················· | ···················· | ···················· | ·······NN·NN········ | 1 |
| I | 3486 hCoV-19/USA/TX-HMH-6224/2020\|EPI_ISL_547325\|2020-0 | ···················· | NNNNNNNNNNNNNNNNNNNN | ···················· | ···················· | ··············N····· | ···N················ | 1 |
| I | 3487 hCoV-19/USA/TX-HMH-6229/2020\|EPI_ISL_547327\|2020-0 | ···················· | ···················· | ···················· | ···················· | NNNNNNNNNNNNNNNNNNNN | NNNNNNNNNNNNNNNNNNNN | 1 |
| I | 3491 hCoV-19/USA/TX-HMH-6240/2020\|EPI_ISL_547334\|2020-0 | ···················· | ·············A······ | ···················· | ···················· | NNNNNNNNNNNNNNNNNNNN | NNNNNNNNNNNNNNNNNNNN | 1 |
| I | 3494 hCoV-19/USA/TX-HMH-6244/2020\|EPI_ISL_547337\|2020-0 | ···················· | ···················· | ···················· | ···················· | NNNNNNNNNNNNNNNNNNNN | NNNNNNNNNNNNNNNNNNNN | 1 |
| I | 3495 hCoV-19/USA/TX-HMH-6246/2020\|EPI_ISL_547339\|2020-0 | ···················· | ·············A······ | ···················· | ···················· | NNNNNNNNNNNNNNNNNNNN | NNNNNNNNNNNNNNNNNNNN | 1 |
| I | 3497 hCoV-19/USA/TX-HMH-6252/2020\|EPI_ISL_547341\|2020-0 | ···················· | ···················· | ···················· | ···················· | NNNNNNNNNNNNNNNNNNNN | NNNNNNNNNNNNNNNNNNNN | 1 |
| I | 3499 hCoV-19/USA/TX-HMH-6255/2020\|EPI_ISL_547343\|2020-0 | ···················· | ···················· | ···················· | ···················· | NNNNNNNNNNNNNNNNNNNN | NNNNNNNNNNNNNNNNNNNN | 1 |
| I | 3501 hCoV-19/USA/TX-HMH-6258/2020\|EPI_ISL_547345\|2020-0 | ···················· | ···················· | ···················· | ···················· | NNNNNNNNNNNNNNNNNNNN | NNNNNNNNNNNNNNNNNNNN | 1 |
| I | 3534 hCoV-19/USA/CA-CZB-4036/2020\|EPI_ISL_548629\|2020-0 | ···················· | ·A·················· | ···················· | ···················· | NNNNNNNNNNNNNNNNNNNN | NNNNNNNNNNNNNNNNNNNN | 1 |
| I | 3577 hCoV-19/USA/CA-CZB-4026/2020\|EPI_ISL_548673\|2020-0 | ···················· | ···················· | ···················· | ···················· | NNNNNNNNNNNNNNNNNNNN | ···················· | 1 |
| I | 3586 hCoV-19/USA/TX-HMH-2864/2020\|EPI_ISL_543834\|2020-0 | ···················· | ···················· | ···················· | ···················· | ········N·N···N····· | NN·····NN··········· | 1 |
| I | 3589 hCoV-19/USA/TX-HMH-2867/2020\|EPI_ISL_543837\|2020-0 | ···················· | ···················· | ···················· | ···················· | NNNNNNNNNNNNNNNNNNNN | NNNNNNNNNNNNNNNNNNNN | 1 |
| I | 3590 hCoV-19/USA/TX-HMH-2868/2020\|EPI_ISL_543838\|2020-0 | ···················· | ···················· | ···················· | ···················· | NNNNNNNNNNNNNNNNNNNN | NNNNNNNNNNNNNNNNNNNN | 1 |
| I | 3607 hCoV-19/USA/TX-HMH-2888/2020\|EPI_ISL_543856\|2020-0 | ···················· | ···················· | ···················· | ···················· | NNNNNNNNNNNNNNNNNNNN | NNNNNNNNNNNNNNNNNNNN | 1 |
| I | 3619 hCoV-19/USA/TX-HMH-2930/2020\|EPI_ISL_543871\|2020-0 | ···················· | ···················· | ···················· | ···················· | NNNNNNNNNNNNNNNNNNNN | NNNNNNNNNNNNNNNNNNNN | 1 |
| I | 3630 hCoV-19/USA/TX-HMH-2958/2020\|EPI_ISL_543892\|2020-0 | ···················· | ···················· | ···················· | ···················· | NNNNNNNNNNNNNNNNNNNN | NNNNNNNNNNNNNNNNNNNN | 1 |
| I | 3631 hCoV-19/USA/TX-HMH-2959/2020\|EPI_ISL_543893\|2020-0 | ···················· | ·A·················· | ···················· | ···················· | N··N···N······NNN··· | NN·N···NN······N···· | 1 |
| I | 3632 hCoV-19/USA/TX-HMH-2960/2020\|EPI_ISL_543894\|2020-0 | ···················· | ···················· | ···················· | ···················· | ··············NN···· | NNNNNNNNN·········NN | 1 |
| I | 3633 hCoV-19/USA/TX-HMH-2961/2020\|EPI_ISL_543895\|2020-0 | ···················· | ···················· | ···················· | ···················· | NNNNNNNNNNNNNNNNNNNN | NNNNNNNNNNNNNNNNNNNN | 1 |
| I | 3639 hCoV-19/USA/TX-HMH-3503/2020\|EPI_ISL_544206\|2020-0 | ···················· | ···················· | ···················· | ···················· | ···················· | NN·N·········NN····· | 1 |
| I | 3642 hCoV-19/USA/TX-HMH-3507/2020\|EPI_ISL_544209\|2020-0 | ···················· | ···················· | ···················· | ···················· | NNNNNNNNNNNNNNNNNNNN | NNNNNNNNNNNNNNNNNNNN | 1 |
| I | 3643 hCoV-19/USA/TX-HMH-3508/2020\|EPI_ISL_544210\|2020-0 | ···················· | ···················· | ···················· | ···················· | NNNNNNNNNNNNNNNNNNNN | NNNNNNNNNNNNNNNNNNNN | 1 |
| I | 3644 hCoV-19/USA/TX-HMH-3511/2020\|EPI_ISL_544211\|2020-0 | ···················· | ······A············· | ···················· | ···················· | NNNNNNNNNNNNNNNNNNNN | NNNNNNNNNNNNNNNNNNNN | 1 |
| I | 3648 hCoV-19/USA/TX-HMH-3520/2020\|EPI_ISL_544219\|2020-0 | ···················· | ···················· | ···················· | ···················· | NNNNNNNNNNNNNNNNNNNN | NNNNNNNNNNNNNNNNNNNN | 1 |
| I | 3651 hCoV-19/USA/TX-HMH-3524/2020\|EPI_ISL_544222\|2020-0 | ···················· | ···················· | ···················· | ···················· | ··············N····· | NN·N···NN··········· | 1 |
| I | 3654 hCoV-19/USA/TX-HMH-3527/2020\|EPI_ISL_544225\|2020-0 | ···················· | ···················· | ···················· | ···················· | NNNNNNNNNNNNNNNNNNNN | NNNNNNNNNNNNNNNNNNNN | 1 |
| I | 3656 hCoV-19/USA/TX-HMH-3529/2020\|EPI_ISL_544227\|2020-0 | ···················· | ···················· | ···················· | ···················· | NNNNNNNNNNNNNNNNNNNN | NNNNNNNNNNNNNNNNNNNN | 1 |
| I | 3658 hCoV-19/USA/TX-HMH-3533/2020\|EPI_ISL_544229\|2020-0 | ···················· | ···················· | ···················· | ···················· | NNNNNNNNNNNNNNNNNNNN | NNNNNNNNNNNNNNNNNNNN | 1 |
| I | 3659 hCoV-19/USA/TX-HMH-3534/2020\|EPI_ISL_544230\|2020-0 | ···················· | ·············A······ | ···················· | ···················· | NNNNNNNNNNNNNNNNNNNN | NNNNNNNNNNNNNNNNNNNN | 1 |
| I | 3660 hCoV-19/USA/TX-HMH-3535/2020\|EPI_ISL_544231\|2020-0 | ···················· | ···················· | ···················· | ···················· | NNNNNNNNNNNNNNNNNNNN | NNNNNNNNNNNNNNNNNNNN | 1 |
| I | 3661 hCoV-19/USA/TX-HMH-3536/2020\|EPI_ISL_544232\|2020-0 | ···················· | ···················· | ···················· | ···················· | NNNNNNNNNNNNNNNNNNNN | NNNNNNNNNNNNNNNNNNNN | 1 |
| I | 3662 hCoV-19/USA/TX-HMH-3538/2020\|EPI_ISL_544233\|2020-0 | ···················· | ···················· | ···················· | ···················· | NNNNNNNNNNNNNNNNNNNN | NNNNNNNNNNNNNNNNNNNN | 1 |
| I | 3663 hCoV-19/USA/TX-HMH-3540/2020\|EPI_ISL_544234\|2020-0 | ···················· | ···················· | ···················· | ···················· | NNNNNNNNNNNNNNNNNNNN | NNNNNNNNNNNNNNNNNNNN | 1 |
| I | 3665 hCoV-19/USA/TX-HMH-4665/2020\|EPI_ISL_544238\|2020-0 | ···················· | ·············A······ | ···················· | ···················· | NNNNNNNNNNNNNNNNNNNN | NNNNNNNNNNNNNNNNNNNN | 1 |
| I | 3672 hCoV-19/USA/TX-HMH-2750/2020\|EPI_ISL_545526\|2020-0 | ···················· | ···················· | ···················· | ···················· | ··N··NNNNN····NN···· | NN·····NN····NN····N | 1 |
| I | 3673 hCoV-19/USA/TX-HMH-2751/2020\|EPI_ISL_545527\|2020-0 | ···················· | ···················· | ···················· | ···················· | NNNNNNNNNNNNNNNNNNNN | NNNNNNNNNNNNNNNNNNNN | 1 |
| I | 3675 hCoV-19/USA/TX-HMH-2753/2020\|EPI_ISL_545529\|2020-0 | ···················· | ···················· | ···················· | ···················· | NNNNNNNNNNNNNNNNNNNN | NNNNNNNNNNNNNNNNNNNN | 1 |
| I | 3679 hCoV-19/USA/TX-HMH-2761/2020\|EPI_ISL_545535\|2020-0 | ···················· | ···················· | ···················· | ···················· | NNNNNNNNNNNNNNNNNNNN | NNNNNNNNNNNNNNNNNNNN | 1 |
| I | 3708 hCoV-19/USA/TX-HMH-2806/2020\|EPI_ISL_545568\|2020-0 | ···················· | ···················· | ···················· | ···················· | NNNNNNNNNNNNNNNNNNNN | NNNNNNNNNNNNNNNNNNNN | 1 |

```
I   3717 hCoV-19/USA/TX-HMH-7733/2020|EPI_ISL_545607|2020-0   ···················· ···················· ···················· ···················· NNNNNNNNNNNNNNNNNNNN NNNNNNNNNNNNNNNNNNNN   1
I   3722 hCoV-19/USA/TX-HMH-7742/2020|EPI_ISL_545612|2020-0   ···················· ···················· ···················· ···················· ··NN·N··NN··N·N····· NNNNN·NNN··········N   1
I   3724 hCoV-19/USA/TX-HMH-6488/2020|EPI_ISL_546032|2020-0   ···················· ·············A······ ···················· ···················· NNNNNNNNNNNNNNNNNNNN NNNNNNNNNNNNNNNNNNNN   1
I   3726 hCoV-19/USA/TX-HMH-6492/2020|EPI_ISL_546035|2020-0   ···················· ···················· ···················· ···················· NNNNNNNNNNNNNNNNNNNN NNNNNNNNNNNNNNNNNNNN   1
I   3746 hCoV-19/USA/TX-HMH-6524/2020|EPI_ISL_546057|2020-0   ···················· ···················· ···················· ···················· NNNNNNNNNNNNNNNNNNNN NNNNNNNNNNNNNNNNNNNN   1
I   3747 hCoV-19/USA/TX-HMH-7565/2020|EPI_ISL_546856|2020-0   ···················· ···················· ···················· ···················· NNNNNNNNNNNNNNNNNNNN NNNNNNNNNNNNNNNNNNNN   1
I   3750 hCoV-19/USA/TX-HMH-7573/2020|EPI_ISL_546863|2020-0   ···················· ···················· ···················· ···················· ·········NNNN·NN···· NNNN···NN·NN·····N·N   1
I   3752 hCoV-19/USA/TX-HMH-7577/2020|EPI_ISL_546865|2020-0   ···················· ···················· ···················· ···················· NNNNNNNNNNNNNNNNNNNN NNNNNNNNNNNNNNNNNNNN   1
I   3755 hCoV-19/USA/TX-HMH-7581/2020|EPI_ISL_546870|2020-0   ···················· ···················· ···················· ···················· NNNNNNNNNNNNNNNNNNNN NNNNNNNNNNNNNNNNNNNN   1
I   3757 hCoV-19/USA/TX-HMH-7586/2020|EPI_ISL_546873|2020-0   ···················· ···················· ···················· ···················· NNNNNNNNNNNNNNNNNNNN NNNNNNNNNNNNNNNNNNNN   1
I   3760 hCoV-19/USA/TX-HMH-7598/2020|EPI_ISL_546881|2020-0   ···················· ···················· ···················· ···················· ········NN····NN··NN NN·N···NNNNN········   1
I   3761 hCoV-19/USA/TX-HMH-7599/2020|EPI_ISL_546882|2020-0   ···················· ···················· ···················· ···················· ············NN······ ··NN···NN···········   1
I   3764 hCoV-19/USA/TX-HMH-7604/2020|EPI_ISL_546886|2020-0   ···················· ···················· ···················· ···················· NNNNNNNNNNNNNNNNNNNN NNNNNNNNNNNNNNNNNNNN   1
I   3765 hCoV-19/USA/TX-HMH-7605/2020|EPI_ISL_546887|2020-0   ···················· ···················· ···················· ···················· NNNNNNNNNNNNNNNNNNNN NNNNNNNNNNNNNNNNNNNN   1
I   3766 hCoV-19/USA/TX-HMH-7606/2020|EPI_ISL_546888|2020-0   ···················· ·············A······ ···················· ···················· NNNNNNNNNNNNNNNNNNNN NNNNNNNNNNNNNNNNNNNN   1
I   3768 hCoV-19/USA/TX-HMH-7612/2020|EPI_ISL_546890|2020-0   ···················· ·············A······ ···················· ···················· NNNNNNNNNNNNNNNNNNNN NNNNNNNNNNNNNNNNNNNN   1
I   3771 hCoV-19/USA/TX-HMH-7624/2020|EPI_ISL_546894|2020-0   ···················· ···················· ···················· ···················· NNNNNNNNNNNNNNNNNNNN NNNNNNNNNNNNNNNNNNNN   1
I   3775 hCoV-19/USA/TX-HMH-7628/2020|EPI_ISL_546898|2020-0   ···················· ···················· ···················· ···················· NNNNNNNNNNNNNNNNNNNN NNNNNNNNNNNNNNNNNNNN   1
I   3776 hCoV-19/USA/TX-HMH-7629/2020|EPI_ISL_546899|2020-0   ···················· ·············A······ ···················· ···················· N·NN·N········NN··NN NNNN···NN····NN·N···   1
I   3785 hCoV-19/USA/TX-HMH-7645/2020|EPI_ISL_546910|2020-0   ···················· ···················· ···················· ···················· NNNNNNNNNNNNNNNNNNNN NNNNNNNNNNNNNNNNNNNN   1
I   3787 hCoV-19/USA/TX-HMH-7649/2020|EPI_ISL_546912|2020-0   ···················· ···················· ···················· ···················· NNNNNNNNNNNNNNNNNNNN NNNNNNNNNNNNNNNNNNNN   1
I   3790 hCoV-19/USA/TX-HMH-7652/2020|EPI_ISL_546916|2020-0   ···················· ···················· ···················· ···················· N··N········NNNN···· NNNN················   1
I   3791 hCoV-19/USA/TX-HMH-7655/2020|EPI_ISL_546918|2020-0   ···················· ·············A······ ···················· ···················· NNNNNNNNNNNNNNNNNNNN NNNNNNNNNNNNNNNNNNNN   1
I   3794 hCoV-19/USA/TX-HMH-7660/2020|EPI_ISL_546922|2020-0   ···················· ···················· ···················· ···················· ··············N····· NN·····NN·····N·····   1
I   3795 hCoV-19/USA/TX-HMH-7665/2020|EPI_ISL_546925|2020-0   ···················· ···················· ···················· ···················· ···················· NN·····NN···········   1
I   3796 hCoV-19/USA/TX-HMH-7669/2020|EPI_ISL_546927|2020-0   ···················· ···················· ···················· ···················· NNNNNNNNNNNNNNNNNNNN NNNNNNNNNNNNNNNNNNNN   1
I   3797 hCoV-19/USA/TX-HMH-7675/2020|EPI_ISL_546931|2020-0   ···················· ···················· ···················· ···················· NNNNNNNNNNNNNNNNNNNN NNNNNNNNNNNNNNNNNNNN   1
I   3799 hCoV-19/USA/TX-HMH-7678/2020|EPI_ISL_546933|2020-0   ···················· ···················· ···················· ···················· NNNNNNNNNNNNNNNNNNNN NNNNNNNNNNNNNNNNNNNN   1
I   3802 hCoV-19/USA/TX-HMH-5675/2020|EPI_ISL_546937|2020-0   ···················· ·A·················· ···················· ···················· ·N·N··········NN···· NN···NNNNN···NN·····   1
I   3803 hCoV-19/USA/TX-HMH-5677/2020|EPI_ISL_546938|2020-0   ···················· ···················· ···················· ···················· NNNNNNNNNNNNNNNNNNNN NNNNNNNNNNNNNNNNNNNN   1
I   3805 hCoV-19/USA/TX-HMH-5680/2020|EPI_ISL_546941|2020-0   ···················· ·············A······ ···················· ···················· NNNNNNNNNNNNNNNNNNNN NNNNNNNNNNNNNNNNNNNN   1
I   3807 hCoV-19/USA/TX-HMH-6283/2020|EPI_ISL_547362|2020-0   ···················· ···················· ···················· ···················· NNNNNNNNNNNNNNNNNNNN NNNNNNNNNNNNNNNNNNNN   1
I   3810 hCoV-19/USA/TX-HMH-6293/2020|EPI_ISL_547368|2020-0   ···················· ···················· ···················· ···················· NNNNNNNNNNNNNNNNNNNN NNNNNNNNNNNNNNNNNNNN   1
I   3811 hCoV-19/USA/TX-HMH-6294/2020|EPI_ISL_547369|2020-0   ···················· ···················· ···················· ···················· NNNNNNNNNNNNNNNNNNNN NNNNNNNNNNNNNNNNNNNN   1
I   3812 hCoV-19/USA/TX-HMH-6297/2020|EPI_ISL_547371|2020-0   ···················· ·············A······ ···················· ···················· ··NNN···N····NN·NN·· NN·N······NN·N····NN   1
I   3815 hCoV-19/USA/TX-HMH-6300/2020|EPI_ISL_547374|2020-0   ···················· ···················· ···················· ···················· ··NNN····NNN··NNN··· NNNN···NN··NNNN····N   1
I   3822 hCoV-19/USA/TX-HMH-6311/2020|EPI_ISL_547382|2020-0   ···················· ···················· ···················· ···················· ···················· N··N···NN··N········   1
I   3848 hCoV-19/USA/TX-HMH-2836/2020|EPI_ISL_543808|2020-0   ···················· ···················· ···················· ···················· NNNNNNNNNNNNNNNNNNNN NNNNNNNNNNNNNNNNNNNN   1
I   3849 hCoV-19/USA/TX-HMH-2838/2020|EPI_ISL_543809|2020-0   ···················· ···················· ···················· ···················· NNNNNNNNNNNNNNNNNNNN NNNNNNNNNNNNNNNNNNNN   1
I   3859 hCoV-19/USA/TX-HMH-2848/2020|EPI_ISL_543819|2020-0   ···················· ···················· ···················· ···················· NNNNNNNNNNNNNNNNNNNN NNNNNNNNNNNNNNNNNNNN   1
I   3860 hCoV-19/USA/TX-HMH-2850/2020|EPI_ISL_543821|2020-0   ···················· ···················· ···················· ···················· NNNNNNNNNNNNNNNNNNNN NNNNNNNNNNNNNNNNNNNN   1
I   3861 hCoV-19/USA/TX-HMH-2851/2020|EPI_ISL_543822|2020-0   ···················· ···················· ···················· ···················· NNNNNNNNNNNNNNNNNNNN NNNNNNNNNNNNNNNNNNNN   1
I   3863 hCoV-19/USA/TX-HMH-2854/2020|EPI_ISL_543825|2020-0   ···················· ···················· ···················· ···················· NNNNNNNNNNNNNNNNNNNN NNNNNNNNNNNNNNNNNNNN   1
I   3865 hCoV-19/USA/TX-HMH-2858/2020|EPI_ISL_543828|2020-0   ···················· ···················· ···················· ···················· NNNNNNNNNNNNNNNNNNNN NNNNNNNNNNNNNNNNNNNN   1
I   3867 hCoV-19/USA/TX-HMH-2860/2020|EPI_ISL_543830|2020-0   ···················· ······A············· ···················· ···················· NNNNNNNNNNNNNNNNNNNN NNNNNNNNNNNNNNNNNNNN   1
I   3877 hCoV-19/USA/TX-HMH-6542/2020|EPI_ISL_546068|2020-0   ···················· ···················· ···················· ···················· NNNNNNNNNNNNNNNNNNNN NNNNNNNNNNNNNNNNNNNN   1
I   3883 hCoV-19/USA/TX-HMH-6548/2020|EPI_ISL_546074|2020-0   ···················· ···················· ···················· ···················· NNNNNNNNNNNNNNNNNNNN NNNNNNNNNNNNNNNNNNNN   1
I   3884 hCoV-19/USA/TX-HMH-655/2020|EPI_ISL_546075|2020-04   ···················· ···················· ···················· ···················· NNNNNNNNNNNNNNNNNNNN ····················   1
I   3885 hCoV-19/USA/TX-HMH-6553/2020|EPI_ISL_546078|2020-0   ···················· ·············A······ ···················· ···················· NN······N·NNN·NN·N·· NN·N···NN··········N   1
I   3903 hCoV-19/USA/TX-HMH-6332/2020|EPI_ISL_547396|2020-0   ···················· ···················· ···················· ···················· ··············N····· NN·····NN···········   1
J   2447 hCoV-19/England/CAMC-947CA6/2020|EPI_ISL_556562|20   ···················· ···················· ···················· NNNNNNNNNNNNNNNNNNNN ···················· ····················   1
J   2892 hCoV-19/England/ALDP-9320FD/2020|EPI_ISL_552940|20   ···················· NNNNNNNNNNNNNNNNNNNN ···················· ···················· ···················· ····················   1
J   3595 hCoV-19/USA/FL-BPHL-1429/2020|EPI_ISL_549211|2020-   ···················· ···················· ···················· ···················· NNNNNNN············· NNN·················   1
J   3605 hCoV-19/USA/FL-BPHL-1439/2020|EPI_ISL_549221|2020-   ···················· ···················· ·········T·········· ···················· NNNNNNNNNNNNNNNNNNNN NNNNNNNNNNNNNNN·····   1
J   3619 hCoV-19/USA/FL-BPHL-1459/2020|EPI_ISL_549241|2020-   ···················· ···················· ···················· ···················· ···················· NNNNN···············   1
J   4155 hCoV-19/USA/FL-BPHL-1407/2020|EPI_ISL_549189|2020-   ···················· ···················· ···················· ···················· NNNNNNNNNNNNNNNNNNNN ····················   1
J   4860 hCoV-19/Wales/PHWC-16B4AC/2020|EPI_ISL_560293|2020   ···················· ···················· ···················· ···················· NNNNNNNNNNNNNNNNNNNN NNNNNNNNNNNNNNNNNNNN   1
J   4863 hCoV-19/Wales/PHWC-16B4E8/2020|EPI_ISL_560297|2020   ···················· ···················· ···················· ···················· NNNN················ ····················   1
J   5092 hCoV-19/Wales/PHWC-16B0E4/2020|EPI_ISL_560244|2020   ···················· ···················· ···················· ···················· NNNNNNNNNNNNNN······ ····················   1
J   5096 hCoV-19/Wales/PHWC-16B13C/2020|EPI_ISL_560248|2020   ···················· ···················· ···················· ···················· NNNNNNNNNNNNNNNNNNNN ····················   1
J   5100 hCoV-19/Wales/PHWC-16B1A5/2020|EPI_ISL_560255|2020   ···················· ···················· ···················· ···················· NNNNNNNNNNNNNNNNNNNN NNNNNNNNNNNNNN······   1
J   5104 hCoV-19/Wales/PHWC-16B1E1/2020|EPI_ISL_560259|2020   ···················· ···················· ···················· ···················· NNNNNNNNNNNNNNNNNNNN NNNNNNNNNNNNNNNNNNNN   1
J   5586 hCoV-19/Wales/PHWC-16A838/2020|EPI_ISL_560111|2020   ···················· ···················· ···················· ···················· NNNN················ ····················   1
J   5587 hCoV-19/Wales/PHWC-16A856/2020|EPI_ISL_560113|2020   ···················· ···················· ···················· ···················· NNNNNNNNNNNNNNNNNNNN ····················   1
J   5590 hCoV-19/Wales/PHWC-16A892/2020|EPI_ISL_560117|2020   ···················· ···················· ···················· ···················· NNNNNNNNNNNNNNNNNNNN NNNNNNNNNNNNNNNNNNNN   1
J   5596 hCoV-19/Wales/PHWC-16A926/2020|EPI_ISL_560126|2020   ···················· ···················· ···················· ···················· NNNNNNNNNNNNNNNNNNNN ····················   1
J   5598 hCoV-19/Wales/PHWC-16A944/2020|EPI_ISL_560128|2020   ···················· ···················· ···················· ···················· NNNNNNNNNNNNNNNNNNNN ····················   1
J   5601 hCoV-19/Wales/PHWC-16A980/2020|EPI_ISL_560132|2020   ···················· ···················· ···················· ···················· NNNNNNNNNNNNNNNNNNNN ····················   1
J   6740 hCoV-19/England/MILK-99DC1A/2020|EPI_ISL_550542|20   ···················· NNNNNNNNNNNNNNNNNNNN ···················· ···················· ···················· ····················   1
K    144 hCoV-19/France/PAC-IHU-0334/2020|EPI_ISL_569461|20   ···················· ···················· ···················· ···················· ···················· NNNNNNNNNNNNNNNNNNNN   1
K    259 hCoV-19/Australia/VIC11223/2020|EPI_ISL_563303|202   ···················· ···················· ···················· ···················· NNNNNNNNNNNNNNNNNNNN ····················   1
K    263 hCoV-19/Australia/VIC11240/2020|EPI_ISL_563307|202   ···················· ···················· ···················· ···················· NNNNNNNNNNNNNNNNNNNN ····················   1
K    278 hCoV-19/Australia/VIC11301/2020|EPI_ISL_563322|202   ···················· ···················· ···················· ···················· NNNN················ ····················   1
K    289 hCoV-19/Australia/VIC11337/2020|EPI_ISL_563334|202   ···················· ···················· ···················· ···················· NNNNNNNNNNNNNNNNNNNN NNNNN··NN·······NNNN   1
K    293 hCoV-19/Australia/VIC11342/2020|EPI_ISL_563338|202   ···················· ···················· ···················· ···················· ··········NNNNNNN··· ····················   1
K    302 hCoV-19/Australia/VIC11371/2020|EPI_ISL_563347|202   ···················· ···················· ···················· ···················· NNNNNNNNNNNNN······· ····················   1
K    309 hCoV-19/Australia/VIC11395/2020|EPI_ISL_563354|202   ···················· ···················· ···················· ···················· NNNNNNNNNNNNNNNNNNNN NNNNNNNNNNNNNNNNNNNN   1
K    311 hCoV-19/Australia/VIC11397/2020|EPI_ISL_563356|202   ···················· ···················· ···················· ···················· NNNNNNNNNNNNNNNNNNNN ····················   1
K    316 hCoV-19/Australia/VIC11412/2020|EPI_ISL_563361|202   ···················· ···················· ···················· ···················· NNNNNNNNNNNNNNNNNNNN ····················   1
K    337 hCoV-19/Australia/VIC11461/2020|EPI_ISL_563382|202   ···················· ···················· ···················· ···················· NNNNNNNNNNNN········ ····················   1
```

| | | | | | | | | | |
|---|---|---|---|---|---|---|---|---|---|
| K | 338 | hCoV-19/Australia/VIC11463/2020\|EPI_ISL_563383\|202 | ···················· | ···················· | ···················· | ···················· | NNNNNNNNNNNNNNNNN··· | ···················· | 1 |
| K | 348 | hCoV-19/Australia/VIC11487/2020\|EPI_ISL_563393\|202 | ···················· | ···················· | ···················· | ···················· | NNNNNNNNNNNN········ | ···················· | 1 |
| K | 540 | hCoV-19/Gambia/3668/2020\|EPI_ISL_560992\|2020-07-30 | ···················· | ···················· | ···················· | ···················· | NNNNNNNNNNNNNNNNNNNN | NNNNNNNNNNNNNNNNNNNN | 1 |
| K | 541 | hCoV-19/Gambia/3671/2020\|EPI_ISL_560994\|2020-07-30 | ···················· | ···················· | ···················· | ···················· | NNNNNNNNNNNNNNNNNNNN | NNNNNNNNNNNNNNNNNNNN | 1 |
| K | 542 | hCoV-19/Gambia/3673/2020\|EPI_ISL_560995\|2020-07-30 | ···················· | ···················· | ···················· | ···················· | NNNNNNNNNNNNNNNNNNNN | NNNNNNNNNNNNNNNNNNNN | 1 |
| K | 563 | hCoV-19/Australia/VIC10979/2020\|EPI_ISL_563178\|202 | ···················· | ···················· | ···················· | ···················· | NNNNNNNNNNNNNNNNNNNN | NNNNNNNNNNNNNNNNNNNN | 1 |
| K | 565 | hCoV-19/Australia/VIC11215/2020\|EPI_ISL_563300\|202 | ···················· | ···················· | ···················· | ···················· | NNNNNNNNNNNNNNNNNNNN | ···················· | 1 |
| K | 814 | hCoV-19/Australia/VIC10998/2020\|EPI_ISL_563190\|202 | ···················· | ···················· | ···················· | ···················· | NNNNNNNNNNNNNNNNNNNN | NNNNNNNNNNNNNNNNNNNN | 1 |
| K | 818 | hCoV-19/Australia/VIC11005/2020\|EPI_ISL_563194\|202 | ···················· | ···················· | ···················· | ···················· | NNNNNNNNNNNNNNNNNNNN | NNNNNNNNNNNNNNNNNNNN | 1 |
| K | 821 | hCoV-19/Australia/VIC11012/2020\|EPI_ISL_563197\|202 | ···················· | ···················· | ···················· | ···················· | NNNNNNNNNNNNNNNNNNNN | NNNN················ | 1 |
| K | 829 | hCoV-19/Australia/VIC11022/2020\|EPI_ISL_563205\|202 | ···················· | ···················· | ···················· | ···················· | NNNNNNNNNNNNNNNNNNNN | NNNNNNNNNNNNNNNNNNNN | 1 |
| K | 837 | hCoV-19/Australia/VIC11034/2020\|EPI_ISL_563213\|202 | ···················· | ···················· | ···················· | ···················· | NNNNNNNNNNNNNNNNNNNN | NNNNNNNNNNNNNNNNNNNN | 1 |
| K | 845 | hCoV-19/Australia/VIC11049/2020\|EPI_ISL_563221\|202 | ···················· | ···················· | ···················· | ···················· | ···················· | NNNNNNNNNNNNNNN····· | 1 |
| K | 851 | hCoV-19/Australia/VIC11060/2020\|EPI_ISL_563227\|202 | ···················· | ···················· | ···················· | ···················· | NNNNNNNNNNNNNNNNNNNN | ···················· | 1 |
| K | 859 | hCoV-19/Australia/VIC11076/2020\|EPI_ISL_563235\|202 | ···················· | ···················· | ···················· | ···················· | NNNNNNNNNNNNNNNNNNNN | NNNNNNNNNNNNNNNNNNNN | 1 |
| K | 862 | hCoV-19/Australia/VIC11082/2020\|EPI_ISL_563238\|202 | ···················· | ···················· | ···················· | ···················· | NNNNNNNNNNNNNNNNNNNN | NNNNNNNNNNNNNNNNNNNN | 1 |
| K | 864 | hCoV-19/Australia/VIC11084/2020\|EPI_ISL_563240\|202 | ···················· | ···················· | ···················· | ···················· | ···················· | NNNNNNNNNNNNNNNNNNNN | 1 |
| K | 884 | hCoV-19/Australia/VIC11129/2020\|EPI_ISL_563260\|202 | ···················· | ···················· | ···················· | ···················· | NNNNNNNNNNNNNNNNNNNN | ···················· | 1 |
| K | 945 | hCoV-19/Australia/VIC7666/2020\|EPI_ISL_564563\|2020 | ···················· | ···················· | ···················· | ···················· | ···················· | NNNNNNNNNNNNNNNNNN·· | 1 |
| K | 983 | hCoV-19/USA/MI-MDHHS-SC21812/2020\|EPI_ISL_565855\|2 | ···················· | ···················· | ···················· | ···················· | ·NNNNNNNNNNNNNNNNNNN | ···················· | 1 |
| K | 1335 | hCoV-19/Australia/VIC10746/2020\|EPI_ISL_563066\|202 | ···················· | ···················· | ···················· | ···················· | NNNNNNNNNNNNNNNNNN·· | NNNN················ | 1 |
| K | 1341 | hCoV-19/Australia/VIC10754/2020\|EPI_ISL_563072\|202 | ···················· | ···················· | ···················· | ···················· | NNNNNNNNNNNNNNNNNNNN | NNNNNNNNNNNNNNNNNNNN | 1 |
| K | 1358 | hCoV-19/Australia/VIC10787/2020\|EPI_ISL_563089\|202 | ···················· | ···················· | ···················· | ···················· | NNNNNNNNN··········· | ···················· | 1 |
| K | 1360 | hCoV-19/Australia/VIC10791/2020\|EPI_ISL_563091\|202 | ···················· | ···················· | ···················· | ···················· | NNNNNNNNNNNNNNNNNNNN | ···················· | 1 |
| K | 1376 | hCoV-19/Australia/VIC10818/2020\|EPI_ISL_563107\|202 | ···················· | ···················· | ···················· | ···················· | NNNNNNNNNNNNNNNNNNNN | NNNNNNNNNNNNNNNNNNNN | 1 |
| K | 1379 | hCoV-19/Australia/VIC10821/2020\|EPI_ISL_563110\|202 | ···················· | ···················· | ···················· | ···················· | NNNNNNNNNNNNNNNNNNNN | NNNNNNNNNNNNNNNNNNNN | 1 |
| K | 1387 | hCoV-19/Australia/VIC10848/2020\|EPI_ISL_563125\|202 | ···················· | ···················· | ···················· | ···················· | NNNNNNNNNNNNNNNNNNNN | ···················· | 1 |
| K | 1389 | hCoV-19/Australia/VIC10856/2020\|EPI_ISL_563127\|202 | ···················· | ···················· | ···················· | ···················· | NNNNNNNNNNNNNNNNNNNN | ···················· | 1 |
| K | 1440 | hCoV-19/Australia/VIC7450/2020\|EPI_ISL_564422\|2020 | ···················· | ·····A·············· | ···················· | ···················· | ···················· | NNNN················ | 1 |
| K | 1478 | hCoV-19/Australia/VIC9763/2020\|EPI_ISL_565699\|2020 | ···················· | ···················· | ···················· | ···················· | NNNNNNNNNNNNNNNNNNNN | NNNNNNNNNNNNNNNNNNNN | 1 |
| K | 1498 | hCoV-19/Australia/VIC9867/2020\|EPI_ISL_565751\|2020 | ···················· | ···················· | ···················· | ···················· | NNNNNNNNNNNNNNNNNNNN | ···················· | 1 |
| K | 1600 | hCoV-19/Australia/VIC10886/2020\|EPI_ISL_563138\|202 | ···················· | ···················· | ···················· | ···················· | NNNNNNNNNNNNNNNNNNNN | ·············N······ | 1 |
| K | 1607 | hCoV-19/Australia/VIC10900/2020\|EPI_ISL_563145\|202 | ···················· | ···················· | ···················· | ···················· | NNNNNNNNNNNNNNNNNNNN | ···················· | 1 |
| K | 1609 | hCoV-19/Australia/VIC10904/2020\|EPI_ISL_563147\|202 | ···················· | ···················· | ···················· | ···················· | ·NNNNNNNNNNNNNNNNNNN | NNNNNNNNNNNNNNNNNN·· | 1 |
| K | 1611 | hCoV-19/Australia/VIC10906/2020\|EPI_ISL_563149\|202 | ···················· | ···················· | ···················· | ···················· | NNNNNN·············· | ···················· | 1 |
| K | 1613 | hCoV-19/Australia/VIC10913/2020\|EPI_ISL_563151\|202 | ···················· | ···················· | ···················· | ···················· | NNNNNNNNNNNNNNNNNNNN | ···················· | 1 |
| K | 1614 | hCoV-19/Australia/VIC10916/2020\|EPI_ISL_563152\|202 | ···················· | ···················· | ···················· | ···················· | NNNNNNNNNNNNNNNNNNNN | ···················· | 1 |
| K | 1651 | hCoV-19/Australia/VIC9765/2020\|EPI_ISL_565701\|2020 | ···················· | ···················· | ···················· | ···················· | NNNNNNNNNNNNNNNNNNNN | ···················· | 1 |
| K | 1653 | hCoV-19/Australia/VIC9767/2020\|EPI_ISL_565703\|2020 | ···················· | ···················· | ···················· | ···················· | NNNNNNNNNNNNNNNNNNNN | NNNNNNNNNNNNNNNNNNNN | 1 |
| K | 1654 | hCoV-19/Australia/VIC9770/2020\|EPI_ISL_565704\|2020 | ···················· | ···················· | ···················· | ···················· | NNNNNNNNNNNNNNNNNNNN | NNNN·············NNN | 1 |
| K | 1656 | hCoV-19/Australia/VIC9774/2020\|EPI_ISL_565706\|2020 | ···················· | ···················· | ···················· | ···················· | ··NNNNNNNNNNNNNNNN·· | ···················· | 1 |
| K | 1665 | hCoV-19/Australia/VIC9786/2020\|EPI_ISL_565715\|2020 | ···················· | ···················· | ···················· | ···················· | NNNNNNNNNNNNNNNNNNNN | NNNNNNNNNNNNNNNNNNNN | 1 |
| K | 1668 | hCoV-19/Australia/VIC9789/2020\|EPI_ISL_565718\|2020 | ···················· | ···················· | ···················· | ···················· | NNNNNNNNNNNNNNNNNNNN | NNNNNNNNNNNNNNNN···· | 1 |
| K | 1675 | hCoV-19/Australia/VIC9801/2020\|EPI_ISL_565725\|2020 | ···················· | ···················· | ···················· | ···················· | NNNNNNNNNNNNNNNNNNNN | ···················· | 1 |
| K | 1676 | hCoV-19/Australia/VIC9802/2020\|EPI_ISL_565726\|2020 | ···················· | ···················· | ···················· | ···················· | NNNNNNNNNNNNNNNNNNNN | ···················· | 1 |
| K | 1678 | hCoV-19/Australia/VIC9806/2020\|EPI_ISL_565728\|2020 | ···················· | ···················· | ···················· | ···················· | NNNNNNNNNNNNNNNNNNNN | NNNNNNNNNNNNNNNNNNNN | 1 |
| K | 1724 | hCoV-19/France/PAC-IHU-0062/2020\|EPI_ISL_569281\|20 | ···················· | ···················· | ···················· | ···················· | NNNNNNNNNNNNNNNNNNNN | ···················· | 1 |
| K | 1744 | hCoV-19/USA/OR-UW-9346/2020\|EPI_ISL_570247\|2020-05 | ···················· | ···················· | ···················· | ···················· | ·····NNNNNN········N | ···················· | 1 |
| K | 1843 | hCoV-19/Australia/VIC719/2020\|EPI_ISL_564270\|2020- | ···················· | ···················· | ···················· | ···················· | NNNNNNNNNNNNNNNNNNNN | ····Y··············· | 1 |
| K | 1903 | hCoV-19/Australia/VIC9517/2020\|EPI_ISL_565583\|2020 | ···················· | ···················· | ···················· | ···················· | NNNNNNNNNNNNNNNNNNNN | NNNNNNNNNNNNNNNNNNNN | 1 |
| K | 1913 | hCoV-19/Australia/VIC9540/2020\|EPI_ISL_565593\|2020 | ···················· | ···················· | ···················· | ···················· | ···················· | NNNNNNNNNNNNNNNNNNNN | 1 |
| K | 1934 | hCoV-19/Australia/VIC9611/2020\|EPI_ISL_565625\|2020 | ···················· | ···················· | ···················· | ···················· | NNNNNNNNNNNNNNNNNNNN | ···················· | 1 |
| K | 1935 | hCoV-19/Australia/VIC9614/2020\|EPI_ISL_565626\|2020 | ···················· | ···················· | ···················· | ···················· | NNNNNNNNNNNNNNNNNNNN | NNNNNNNNNNNNNNNNNN·· | 1 |
| K | 1936 | hCoV-19/Australia/VIC9615/2020\|EPI_ISL_565627\|2020 | ···················· | ···················· | ···················· | ···················· | NNNNNNNNNNNN········ | ···················· | 1 |
| K | 1938 | hCoV-19/Australia/VIC9619/2020\|EPI_ISL_565629\|2020 | ···················· | ···················· | ···················· | ···················· | NNNNNNNNNNNNNNNNNNNN | NNNNNNNNNNNNNNNNNNNN | 1 |
| K | 1939 | hCoV-19/Australia/VIC9621/2020\|EPI_ISL_565630\|2020 | ···················· | ···················· | ···················· | ···················· | NNNNNNNNNNNNNNNNNNNN | NNNNNNNNNNNNNNNNNNNN | 1 |
| K | 1940 | hCoV-19/Australia/VIC9624/2020\|EPI_ISL_565631\|2020 | ···················· | ···················· | ···················· | ···················· | ··NNNNNNNNNNNNNNNNN· | NN·················· | 1 |
| K | 2055 | hCoV-19/Australia/VIC10639/2020\|EPI_ISL_563006\|202 | ···················· | ···················· | ···················· | ···················· | NNNNNNNNNNNNNNNNNNNN | NNNNNNNNNNNNNNNNNNNN | 1 |
| K | 2072 | hCoV-19/Australia/VIC10665/2020\|EPI_ISL_563025\|202 | ···················· | ···················· | ···················· | ···················· | NNNNNNNNNNNNNNNNNNNN | NNNNNNNNNNNNNNNNNNNN | 1 |
| K | 2140 | hCoV-19/Australia/VIC9714/2020\|EPI_ISL_565668\|2020 | ···················· | ···················· | ···················· | ···················· | NNNNNNNNNNNNNNNNNNNN | NNNNNNNNNNNNNNNNNNNN | 1 |
| K | 2141 | hCoV-19/Australia/VIC9715/2020\|EPI_ISL_565669\|2020 | ···················· | ···················· | ···················· | ···················· | NNNNNNNNNNNNNNNNNNNN | NNNNNNNNNNNNNNNNNNNN | 1 |
| K | 2144 | hCoV-19/Australia/VIC9720/2020\|EPI_ISL_565672\|2020 | ···················· | ···················· | ···················· | ···················· | NNNNNNNNNNNNNNNN···· | ···················· | 1 |
| K | 2152 | hCoV-19/Australia/VIC9732/2020\|EPI_ISL_565680\|2020 | ···················· | ···················· | ···················· | ···················· | NNNNNNNNNNNNNNNNNNNN | ···················· | 1 |
| K | 2153 | hCoV-19/Australia/VIC9733/2020\|EPI_ISL_565681\|2020 | ···················· | ···················· | ···················· | ···················· | NNNNNNNNNNNNNNNNNNNN | NNNNNNNNNNNNNNNNNNNN | 1 |
| K | 2155 | hCoV-19/Australia/VIC9736/2020\|EPI_ISL_565683\|2020 | ···················· | ···················· | ···················· | ···················· | ··NNNNNNNNNNNNNNNNNN | NNNNNNNNNNNNNNNNNNNN | 1 |
| K | 2158 | hCoV-19/Australia/VIC9740/2020\|EPI_ISL_565686\|2020 | ···················· | ···················· | ···················· | ···················· | NNNNNNNNNNNNNNNNNNNN | NNNNNNNNNNNNNNNNNNNN | 1 |
| K | 2159 | hCoV-19/Australia/VIC9744/2020\|EPI_ISL_565687\|2020 | ···················· | ···················· | ···················· | ···················· | ···················· | NNNNNNNNNNNNNNNNNNNN | 1 |
| K | 2160 | hCoV-19/Australia/VIC9745/2020\|EPI_ISL_565688\|2020 | ···················· | ···················· | ···················· | ···················· | NNNNNNNNNNNNNNNNNNNN | NNNNNNNNNNNNNNNNNNNN | 1 |
| K | 2170 | hCoV-19/Australia/VIC9762/2020\|EPI_ISL_565698\|2020 | ···················· | ···················· | ···················· | ···················· | NNNNNNNNNNNNNNNNNNNN | NNNNNNNNN··········· | 1 |
| K | 2228 | hCoV-19/France/PAC-IHU-1468/2020\|EPI_ISL_569173\|20 | ···················· | ···················· | ···················· | ···················· | ···················· | NNNNNNNNNNNNNNNNNNNN | 1 |
| K | 2314 | hCoV-19/Australia/VIC13387/2020\|EPI_ISL_563709\|202 | ···················· | ···················· | ···················· | ···················· | ··NNNNNNNNNNNNNNN··· | ···················· | 1 |
| K | 2561 | hCoV-19/Gambia/GC19-2707/2020\|EPI_ISL_561134\|2020- | ···················· | ···················· | ···················· | ···················· | NNNNNNNNNNNNNNNNNNNN | NNNNNNNNNNNNNNNNNNNN | 1 |
| K | 2563 | hCoV-19/Gambia/GC19-2730/2020\|EPI_ISL_561136\|2020- | ···················· | ···················· | ···················· | ···················· | NNNNNNNNNNNNNNNNNNNN | NNNNNNNNNNNNNNNNNNNN | 1 |
| K | 2586 | hCoV-19/Gambia/GC19-3085/2020\|EPI_ISL_561163\|2020- | ···················· | ···················· | ···················· | ···················· | NNNNNNNNNNNNNNNNNNNN | NNNNNNNNNNNNNNNNNNNN | 1 |
| K | 2592 | hCoV-19/Gambia/GC19-3135/2020\|EPI_ISL_561169\|2020- | ···················· | ···················· | ···················· | ···················· | NNNNNNNNNNNNNNNNNNNN | NNNNNNNNNNNNNNNNNNNN | 1 |
| K | 2594 | hCoV-19/Gambia/GC19-3146/2020\|EPI_ISL_561171\|2020- | ···················· | ···················· | ···················· | ···················· | NNNNNNNNNNNNNNNNNNNN | NNNNNNNNNNNNNNNNNNNN | 1 |
| K | 2601 | hCoV-19/Gambia/GC19-3432/2020\|EPI_ISL_561179\|2020- | ···················· | ···················· | ···················· | ···················· | NNNNNNNNNNNNNNNNNNNN | NNNNNNNNNNNNNNNNNNNN | 1 |
| K | 2607 | hCoV-19/Gambia/GC19-3504/2020\|EPI_ISL_561187\|2020- | ···················· | ···················· | ···················· | ···················· | NNNNNNNNNNNNNNNNNNNN | NNNNNNNNNNNNNNNNNNNN | 1 |
| K | 2636 | hCoV-19/Australia/VIC13429/2020\|EPI_ISL_563728\|202 | ···················· | ···················· | ···················· | ···················· | NNNNNNNNNNNNNNNNNNNN | NNNN················ | 1 |
| K | 2647 | hCoV-19/Australia/VIC13654/2020\|EPI_ISL_563797\|202 | ···················· | ···················· | ···················· | ···················· | NNNNNNNNNNNN·NNNNNNN | ···················· | 1 |

K 2712 hCoV-19/USA/FL-BPHL-1503/2020|EPI_ISL_568601|2020- ···················· ···················· ···················· ···················· ·············NNNNN·· ···················· 1
K 2836 hCoV-19/Gambia/GC19-2632/2020|EPI_ISL_561124|2020- ···················· ···················· ···················· ···················· NNNNNNNNNNNNNNNNNNNN NNNNNNNNNNNNNNNNNNNN 1
K 2852 hCoV-19/Australia/VIC13463/2020|EPI_ISL_563738|202 ···················· ···················· ···················· ···················· NNNNNNNNNNNNNNNNNNNN ···················· 1
K 3082 hCoV-19/Gambia/GC19-3166/2020|EPI_ISL_561044|2020- ···················· ···················· ···················· ···················· NNNNNNNNNNNNNNNNNNNN NNNNNNNNNNNNNNNNNNNN 1
K 3083 hCoV-19/Gambia/GC19-3192/2020|EPI_ISL_561046|2020- ···················· ···················· ···················· ···················· NNNNNNNNNNNNNNNNNNNN NNNNNNNNNNNNNNNNNNNN 1
K 3084 hCoV-19/Gambia/GC19-3193/2020|EPI_ISL_561047|2020- ···················· ···················· ···················· ···················· NNNNNNNNNNNNNNNNNNNN NNNNNNNNNNNNNNNNNNNN 1
K 3085 hCoV-19/Gambia/GC19-3208/2020|EPI_ISL_561048|2020- ···················· ···················· ···················· ···················· NNNNNNNNNNNNNNNNNNNN NNNNNNNNNNNNNNNNNNNN 1
K 3086 hCoV-19/Gambia/GC19-3209/2020|EPI_ISL_561049|2020- ···················· ···················· ···················· ···················· NNNNNNNNNNNNNNNNNNNN NNNNNNNNNNNNNNNNNNNN 1
K 3087 hCoV-19/Gambia/GC19-3214/2020|EPI_ISL_561051|2020- ···················· ···················· ···················· ···················· NNNNNNNNNNNNNNNNNNNN NNNNNNNNNNNNNNNNNNNN 1
K 3089 hCoV-19/Gambia/GC19-3246/2020|EPI_ISL_561053|2020- ···················· ···················· ···················· ···················· NNNNNNNNNNNNNNNNNNNN NNNNNNNNNNNNNNNNNNNN 1
K 3090 hCoV-19/Gambia/GC19-3255/2020|EPI_ISL_561054|2020- ···················· ···················· ···················· ···················· NNNNNNNNNNNNNNNNNNNN NNNNNNNNNNNNNNNNNNNN 1
K 3091 hCoV-19/Gambia/GC19-3266/2020|EPI_ISL_561055|2020- ···················· ···················· ···················· ···················· NNNNNNNNNNNNNNNNNNNN NNNNNNNNNNNNNNNNNNNN 1
K 3092 hCoV-19/Gambia/GC19-3267/2020|EPI_ISL_561056|2020- ···················· ···················· ···················· ···················· NNNNNNNNNNNNNNNNNNNN NNNNNNNNNNNNNNNNNNNN 1
K 3093 hCoV-19/Gambia/GC19-3278/2020|EPI_ISL_561058|2020- ···················· ···················· ···················· ···················· NNNNNNNNNNNNNNNNNNNN NNNNNNNNNNNNNNNNNNNN 1
K 3094 hCoV-19/Gambia/GC19-3301/2020|EPI_ISL_561059|2020- ···················· ···················· ···················· ···················· NNNNNNNNNNNNNNNNNNNN NNNNNNNNNNNNNNNNNNNN 1
K 3096 hCoV-19/Gambia/GC19-3323/2020|EPI_ISL_561061|2020- ···················· ···················· ···················· ···················· NNNNNNNNNNNNNNNNNNNN NNNNNNNNNNNNNNNNNNNN 1
K 3098 hCoV-19/Gambia/GC19-3365/2020|EPI_ISL_561063|2020- ···················· ···················· ···················· ···················· NNNNNNNNNNNNNNNNNNNN NNNNNNNNNNNNNNNNNNNN 1
K 3099 hCoV-19/Gambia/GC19-3373/2020|EPI_ISL_561064|2020- ···················· ···················· ···················· ···················· NNNNNNNNNNNNNNNNNNNN NNNNNNNNNNNNNNNNNNNN 1
K 3100 hCoV-19/Gambia/GC19-3374/2020|EPI_ISL_561065|2020- ···················· ···················· ···················· ···················· NNNNNNNNNNNNNNNNNNNN NNNNNNNNNNNNNNNNNNNN 1
K 3101 hCoV-19/Gambia/GC19-3375/2020|EPI_ISL_561066|2020- ···················· ···················· ···················· ···················· NNNNNNNNNNNNNNNNNNNN NNNNNNNNNNNNNNNNNNNN 1
K 3102 hCoV-19/Gambia/GC19-3380/2020|EPI_ISL_561067|2020- ···················· ···················· ···················· ···················· NNNNNNNNNNNNNNNNNNNN NNNNNNNNNNNNNNNNNNNN 1
K 3119 hCoV-19/Gambia/GC19-3812/2020|EPI_ISL_561086|2020- ···················· ···················· ···················· ···················· NNNNNNNNNNNNNNNNNNNN ···················· 1
K 3122 hCoV-19/Gambia/GC19-3816/2020|EPI_ISL_561089|2020- ···················· ···················· ···················· ···················· NNNNNNNNNNNNNNNNNNNN NNNNNNNNNNNNNNNNNNNN 1
K 3126 hCoV-19/Gambia/GC19-3823/2020|EPI_ISL_561094|2020- ···················· ···················· ···················· ···················· NNNNNNNNNNNNNNNNNNNN NNNNNNNNNNNNNNNNNNNN 1
K 3154 hCoV-19/Australia/VIC13370/2020|EPI_ISL_563696|202 ···················· ···················· ···················· ···················· NNNNNNNNNNNNNNNNNNNN ···················· 1
K 3340 hCoV-19/Australia/VIC12888/2020|EPI_ISL_563604|202 ···················· ···················· ···················· ···················· NNNNNNNNNNNNNNNNNNNN ···················· 1
K 3344 hCoV-19/Australia/VIC1293/2020|EPI_ISL_563609|2020 ···················· ···················· ···················· ···················· NNNNNNNNNNNNNNNNNNNN NNNNNNNNNNNNNNNNNNNN 1
K 3406 hCoV-19/Australia/VIC13320/2020|EPI_ISL_563671|202 ···················· ···················· ···················· ···················· NNNNNNNNNNNNNNNNNNNN ···················· 1
K 3610 hCoV-19/Australia/VIC1278/2020|EPI_ISL_563594|2020 ···················· ···················· ···················· ···················· NNNNNNNNNNNNNNNNNNNN NNNNNNNNNNNNNNNNNNNN 1
K 3841 hCoV-19/Australia/VIC11973/2020|EPI_ISL_563500|202 ···················· ···················· ···················· ···················· NNNNNNNNNNNNNNNNNNNN ···················· 1
K 3865 hCoV-19/Australia/VIC12114/2020|EPI_ISL_563524|202 ···················· ···················· ···················· ···················· ················W··· NNNNNNNNN·····NNNN·· 1
K 3866 hCoV-19/Australia/VIC12122/2020|EPI_ISL_563525|202 ···················· ···················· ···················· ···················· NNNNNNNNNNNNNNNNNNNN ···················· 1
K 3868 hCoV-19/Australia/VIC12129/2020|EPI_ISL_563527|202 ···················· ···················· ···················· ···················· NNNNNNNNNNNNNNNNNNNN ·········Y·········· 1
K 3869 hCoV-19/Australia/VIC12137/2020|EPI_ISL_563528|202 ···················· ···················· ···················· ···················· NNNNNNNNNNNN········ ·····M·············· 1
K 3878 hCoV-19/Australia/VIC12195/2020|EPI_ISL_563537|202 ···················· ···················· ···················· ···················· NNNNNNNNNNNNNNN····· ···················· 1
K 3882 hCoV-19/Australia/VIC12214/2020|EPI_ISL_563541|202 ···················· ···················· ···················· ···················· NNNN················ ···················· 1
K 3885 hCoV-19/Australia/VIC12228/2020|EPI_ISL_563544|202 ···················· ···················· ···················· ···················· NNNNNNNNNNNNNNNNNNNN ···················· 1
K 3890 hCoV-19/Australia/VIC12246/2020|EPI_ISL_563549|202 ···················· ···················· ···················· ···················· NNNN··R·NNNNNNNNNNNN ···················· 1
K 3892 hCoV-19/Australia/VIC1232/2020|EPI_ISL_563551|2020 ···················· ···················· ···················· ···················· NNNNNNNNNNNNNNNNNNNN NNNNNNNNNNNNNNNNNNNN 1
K 3907 hCoV-19/Australia/VIC12772/2020|EPI_ISL_563591|202 ···················· ···················· ···················· ···················· NNNNNNNNNNNN········ ···················· 1
K 3912 hCoV-19/Australia/VIC12816/2020|EPI_ISL_563598|202 ···················· ···················· ···················· ···················· ·········NNNNNNNNNNN ··NNNNNNNNNNNNNNNNNN 1
K 3962 hCoV-19/Australia/VIC8100/2020|EPI_ISL_564848|2020 ···················· ···················· ···················· ···················· NNNNNNNNNNNNNNNNNNNN NNNNNNNNNNNNNNNNNNNN 1
K 4097 hCoV-19/Australia/VIC11614/2020|EPI_ISL_563433|202 ···················· ···················· ···················· ···················· NNNNNNNNNNNNNNNNNNNN ···················· 1
K 4099 hCoV-19/Australia/VIC11617/2020|EPI_ISL_563435|202 ···················· ···················· ···················· ···················· NNNNNNNNNNNNNNNNNNNN NNNNNNNNNNNNNNNNNNNN 1
K 4100 hCoV-19/Australia/VIC11625/2020|EPI_ISL_563436|202 ···················· ···················· ···················· ···················· NNNNNNNNNNNN········ NNNNNNNNNNNNNNNNNNNN 1
K 4108 hCoV-19/Australia/VIC11641/2020|EPI_ISL_563444|202 ···················· ···················· ···················· ···················· NNNNNNNNNNNNNNNNNNNN NNNNNNNNNNNNNNNNNNNN 1
K 4109 hCoV-19/Australia/VIC11648/2020|EPI_ISL_563445|202 ···················· ···················· ···················· ···················· NNNNNNNNNNNNNNNNNNNN ···················· 1
K 4114 hCoV-19/Australia/VIC11662/2020|EPI_ISL_563450|202 ···················· ···················· ···················· ···················· NNNNNNNNNNNNNN······ ···················· 1
K 4120 hCoV-19/Australia/VIC11678/2020|EPI_ISL_563456|202 ···················· ···················· ···················· ···················· NNNNNNNNNNNNNNNNNNNN NNNNNNNNNNNNNNNNNNNN 1
K 4125 hCoV-19/Australia/VIC11707/2020|EPI_ISL_563466|202 ···················· ···················· ···················· ···················· NNNNNNNNNNNNNNNNNNNN ···················· 1
K 4362 hCoV-19/Australia/VIC10132/2020|EPI_ISL_562802|202 ···················· ···················· ···················· ···················· NNNNNNNNNNNNNNNNN··· NNNNNNNNNNNNNNN····· 1
K 4379 hCoV-19/Australia/VIC10189/2020|EPI_ISL_562819|202 ···················· ···················· ···················· ···················· NNNNNNNNNNNNNNNNNNNN ···················· 1
K 4380 hCoV-19/Australia/VIC10191/2020|EPI_ISL_562820|202 ···················· ···················· ···················· NNNNNNNNNNNNNNNNNNNN NNNNNNNNNNNN········ ···················· 1
K 4385 hCoV-19/Australia/VIC10215/2020|EPI_ISL_562825|202 ···················· ···················· ···················· ···················· NNNNNNNNNNNNNNNNNNNN ···················· 1
K 4387 hCoV-19/Australia/VIC10218/2020|EPI_ISL_562827|202 ···················· ···················· ···················· ···················· NNNNNNNNNNNNNNNNNNNN ···················· 1
K 4393 hCoV-19/Australia/VIC10232/2020|EPI_ISL_562833|202 ···················· ···················· ···················· ···················· NNNNNNNNNNNNN······· ···················· 1
K 4410 hCoV-19/Australia/VIC10284/2020|EPI_ISL_562852|202 ···················· ···················· ···················· ···················· NNNNNNNNNNNNNNNNNNNN NNNNNNNNNNNNNNNNNNNN 1
K 4412 hCoV-19/Australia/VIC10288/2020|EPI_ISL_562854|202 ···················· ···················· ···················· ···················· NNNNNNNNNNNNNNNNNNNN ···················· 1
K 4418 hCoV-19/Australia/VIC10302/2020|EPI_ISL_562860|202 ···················· ···················· ···················· ···················· NN··NNNNNNNNNNNNNNNN NNNNNNNNNNNNNNNNNNNN 1
K 4422 hCoV-19/Australia/VIC11516/2020|EPI_ISL_563403|202 ···················· ···················· ···················· ···················· NNNNNNNNNNNNNN······ NNNNNNNNNNNNNNN····· 1
K 4428 hCoV-19/Australia/VIC11548/2020|EPI_ISL_563409|202 ···················· ···················· ···················· ···················· NNNNNNNNNNNNNNNNNNNN NNNNNNNNNNNNNNNNNNNN 1
K 4429 hCoV-19/Australia/VIC11549/2020|EPI_ISL_563410|202 ···················· ···················· ···················· ···················· NNNNNNNNNNNNNNNNNNNN ···················· 1
K 4434 hCoV-19/Australia/VIC11560/2020|EPI_ISL_563415|202 ···················· ···················· ···················· ···················· NNNNNNNNNNNNNNNNNNNN ···················· 1
K 4436 hCoV-19/Australia/VIC11573/2020|EPI_ISL_563417|202 ···················· ···················· ···················· ···················· NNNNNNNNNNNNNNNNNNNN ···················· 1
K 4437 hCoV-19/Australia/VIC11575/2020|EPI_ISL_563418|202 ···················· ···················· ···················· ···················· NNNNNNNNNNNNNNNNN··· ···················· 1
K 4440 hCoV-19/Australia/VIC11583/2020|EPI_ISL_563421|202 ···················· ···················· ···················· ···················· NNNNNNNNNNNNNNNN···· ···················· 1
K 4441 hCoV-19/Australia/VIC11585/2020|EPI_ISL_563422|202 ···················· ···················· ···················· ···················· NNNNNNNNNNNNNNNNNNNN ···················· 1
K 4442 hCoV-19/Australia/VIC11590/2020|EPI_ISL_563423|202 ···················· ···················· ···················· ···················· NNNNNNNNNNNNNNNNNNNN NNNNNNNNNNNNNNNNNNNN 1
K 4450 hCoV-19/Australia/VIC11611/2020|EPI_ISL_563431|202 ···················· ···················· ···················· ···················· ····NNNNNNNN········ NNNNNNNNNNNNNNNN···· 1
K 4456 hCoV-19/Australia/VIC11705/2020|EPI_ISL_563465|202 ···················· ···················· ···················· ···················· NNNNNNNNNNNNNNNNNNNN NNNNNNNNN·····NNNNNN 1
K 4473 hCoV-19/Australia/VIC11767/2020|EPI_ISL_563493|202 ···················· ···················· ···················· ···················· NNNNNNNNNNNNNNNN···· NNNNNNNNN··········· 1
K 4474 hCoV-19/Australia/VIC11773/2020|EPI_ISL_563494|202 ···················· ···················· ···················· ···················· NNNNNNNNNNNNNNNNNNNN ···················· 1
K 4586 hCoV-19/Australia/VIC8570/2020|EPI_ISL_565096|2020 ···················· ···················· ···················· ···················· NNNNNNNNNNNNNNN····· ···················· 1
K 4661 hCoV-19/Australia/VIC8490/2020|EPI_ISL_565050|2020 ···················· ······A············· ···················· ···················· NNNNNNNNNNNNNNNNNNNN NNNNNNNNNNNNNNNNNNNN 1
K 4879 hCoV-19/Australia/VIC10384/2020|EPI_ISL_562733|202 ···················· ···················· ···················· ···················· NNNNNNNNNNNN········ ···················· 1
K 4894 hCoV-19/Australia/VIC10321/2020|EPI_ISL_562869|202 ···················· ···················· ···················· ···················· NNNNNNNNNNNN········ ···················· 1
K 4901 hCoV-19/Australia/VIC10336/2020|EPI_ISL_562876|202 ···················· ······A············· ···················· ···················· NNNNNNNNNNNNNNNNNNNN ···················· 1
K 4902 hCoV-19/Australia/VIC10337/2020|EPI_ISL_562877|202 ···················· ···················· ···················· ···················· NNNNNNNNNNNNNNNNNNNN NNNNNNNNNNNNNNNNNNNN 1
K 4905 hCoV-19/Australia/VIC10343/2020|EPI_ISL_562880|202 ···················· ···················· ···················· ···················· NNNNNNNNNNNNNNNNNNNN NNNNNNNNNNNNNNNNNNNN 1
K 4907 hCoV-19/Australia/VIC10350/2020|EPI_ISL_562882|202 ···················· ···················· ···················· ···················· NNNNNNNNNNNNNNNNNNNN NNNNNNNNN··········· 1

| | | | | | | | | |
|---|---|---|---|---|---|---|---|---|
| K | 4914 hCoV-19/Australia/VIC10361/2020\|EPI_ISL_562889\|202 | ···················· | ···················· | ···················· | ···················· | NNNNNNNNNNNNNNNNNNNN | NNNNNNNNNNNNNNNNNNNN | 1 |
| K | 4915 hCoV-19/Australia/VIC10362/2020\|EPI_ISL_562890\|202 | ···················· | ···················· | ···················· | ···················· | NNNNNNNNNNNNNNNNNNNN | NNNNNNNNNNNNNNNNNNNN | 1 |
| K | 4918 hCoV-19/Australia/VIC10378/2020\|EPI_ISL_562893\|202 | ···················· | ···················· | ···················· | ···················· | NNNNNNNNNNNNNNNNNNNN | NNNNN···········W··· | 1 |
| K | 4920 hCoV-19/Australia/VIC10381/2020\|EPI_ISL_562895\|202 | ···················· | ···················· | ···················· | ···················· | NNNNNNNNNNNNNNNNNNNN | ···················· | 1 |
| K | 5121 hCoV-19/Australia/VIC14035/2020\|EPI_ISL_562740\|202 | ···················· | ···················· | ···················· | ···················· | NNNNNNNNNNNNNNNNNNNN | NNNNNNNNNNNNNNNNNNNN | 1 |
| K | 5122 hCoV-19/Australia/VIC14070/2020\|EPI_ISL_562741\|202 | ···················· | ···················· | ···················· | ···················· | NNNNNNNNNNNNNNNNNNNN | NNNNNNNNNNNNNNNNNNNN | 1 |
| K | 5131 hCoV-19/Australia/VIC9901/2020\|EPI_ISL_562750\|2020 | ···················· | ···················· | ···················· | ···················· | NNNNNNNNNNNNNNNNNNNN | NNNNNNNNNNNNNNNN···· | 1 |
| K | 5176 hCoV-19/Australia/VIC13969/2020\|EPI_ISL_563902\|202 | ···················· | ···················· | ···················· | ···················· | NNNNNNNNNNNNNNNNNNNN | NNNNNNNNNNNNNNNNNNNN | 1 |
| K | 5179 hCoV-19/Australia/VIC13977/2020\|EPI_ISL_563905\|202 | ···················· | ···················· | ···················· | ···················· | NNNNNNNNNNNNNNNNNNNN | ···················· | 1 |
| K | 5180 hCoV-19/Australia/VIC13980/2020\|EPI_ISL_563906\|202 | ···················· | ···················· | ···················· | ···················· | NNNNNNNNNNNNNNNNNNNN | NNNNNNNNNNNNNNN····· | 1 |
| K | 5181 hCoV-19/Australia/VIC13981/2020\|EPI_ISL_563907\|202 | ···················· | ···················· | ···················· | ···················· | NNNNNNNNNNNNNNNN···· | ···················· | 1 |
| K | 5182 hCoV-19/Australia/VIC13982/2020\|EPI_ISL_563908\|202 | ···················· | ···················· | ···················· | ···················· | NNNNNNNNNNNNNNNNNNNN | ···················· | 1 |
| K | 5185 hCoV-19/Australia/VIC14006/2020\|EPI_ISL_563922\|202 | ···················· | ···················· | ···················· | ···················· | NNNNNNNNNNNNNNNNN··· | ···················· | 1 |
| K | 5187 hCoV-19/Australia/VIC1401/2020\|EPI_ISL_563925\|2020 | ···················· | ···················· | ···················· | ···················· | NNNNNNNNNNNNNNNNNNNN | NNNNNNNNNNNNNNNNNNNN | 1 |
| K | 5350 hCoV-19/France/PAC-IHU-0686/2020\|EPI_ISL_568934\|20 | ···················· | ···················· | ···················· | NNNNNN·············· | ···················· | ···················· | 1 |
| K | 5387 hCoV-19/Australia/VIC10501/2020\|EPI_ISL_562601\|202 | ···················· | ···················· | ···················· | ···················· | NNNNNNNNNNNNNNNNNNNN | ·····NNNNNNNNNN····· | 1 |
| K | 5398 hCoV-19/Australia/VIC11425/2020\|EPI_ISL_562612\|202 | ···················· | ···················· | ···················· | ···················· | NNNNNNNNNNNNNNNNNNNN | ···················· | 1 |
| K | 5400 hCoV-19/Australia/VIC11439/2020\|EPI_ISL_562614\|202 | ···················· | ···················· | ···················· | ···················· | NNNNNNNNNNNNNNNNNNNN | ···················· | 1 |
| K | 5401 hCoV-19/Australia/VIC12465/2020\|EPI_ISL_562615\|202 | ···················· | ···················· | ···················· | ···················· | NNNNNNNNNNNNNNNNNNNN | NNNNNNNNNNNNNNNNNNNN | 1 |
| K | 5406 hCoV-19/Australia/VIC13664/2020\|EPI_ISL_562620\|202 | ···················· | ···················· | ···················· | ···················· | NNNNNNNNNNNNNNNNNNNN | NNNNNNNNNNNNNNNNNNNN | 1 |
| K | 5408 hCoV-19/Australia/VIC14039/2020\|EPI_ISL_562622\|202 | ···················· | ···················· | ···················· | ···················· | NNNNNNNNNNNNNNNNNNNN | NNNNNNNNNNNNNNNNNNNN | 1 |
| K | 5424 hCoV-19/Australia/VIC10260/2020\|EPI_ISL_562638\|202 | ···················· | ···················· | ···················· | ···················· | NNNNNNNNNNNNNNNNNNNN | ···················· | 1 |
| K | 5427 hCoV-19/Australia/VIC13776/2020\|EPI_ISL_562653\|202 | ···················· | ···················· | ···················· | ···················· | NNNNNNNNNNNNNNNNNNNN | ···················· | 1 |
| K | 5428 hCoV-19/Australia/VIC13925/2020\|EPI_ISL_562654\|202 | ···················· | ···················· | ···················· | ···················· | NNNNNNNNNNNNN······· | NNNN················ | 1 |
| K | 5431 hCoV-19/Australia/VIC13988/2020\|EPI_ISL_563911\|202 | ···················· | ···················· | ···················· | ···················· | NNNNNNNNNNNNNNNNNNNN | ···················· | 1 |
| K | 5433 hCoV-19/Australia/VIC13990/2020\|EPI_ISL_563913\|202 | ···················· | ···················· | ···················· | ···················· | NNNNNNNNNNNNNNNNNNN· | NNNN················ | 1 |
| K | 5435 hCoV-19/Australia/VIC13993/2020\|EPI_ISL_563915\|202 | ···················· | ···················· | ···················· | ···················· | NNNNNNNNNNNNN······· | ···················· | 1 |
| K | 5438 hCoV-19/Australia/VIC13999/2020\|EPI_ISL_563918\|202 | ···················· | ···················· | ···················· | ···················· | NNNNNNNNNNNNNNNNNNNN | ···················· | 1 |
| K | 5443 hCoV-19/Australia/VIC14017/2020\|EPI_ISL_563931\|202 | ···················· | ···················· | ···················· | ···················· | NNNNNNNNNNNNNNNNNNNN | ···················· | 1 |
| K | 5444 hCoV-19/Australia/VIC14018/2020\|EPI_ISL_563932\|202 | ···················· | ···················· | ···················· | ···················· | ···················· | NNNN················ | 1 |
| K | 5445 hCoV-19/Australia/VIC14020/2020\|EPI_ISL_563933\|202 | ···················· | ···················· | ···················· | ···················· | NNNNNNNNNNNN········ | ···················· | 1 |
| K | 5447 hCoV-19/Australia/VIC14025/2020\|EPI_ISL_563935\|202 | ···················· | ···················· | ···················· | ···················· | NNNNNNNNNNNNNNNNNNNN | ···················· | 1 |
| K | 5448 hCoV-19/Australia/VIC14026/2020\|EPI_ISL_563936\|202 | ···················· | ···················· | ···················· | ···················· | NNNNNNNNNNNNNNNNNNNN | NNNNNNNNNNNNNNNNNNNN | 1 |
| K | 5449 hCoV-19/Australia/VIC14027/2020\|EPI_ISL_563937\|202 | ···················· | ···················· | ···················· | ···················· | NNNNNNNNNNNNNNNNNNNN | NNNN················ | 1 |
| K | 5451 hCoV-19/Australia/VIC14034/2020\|EPI_ISL_563939\|202 | ···················· | ···················· | ···················· | ···················· | ···················· | NNNNNNNNNN·········· | 1 |
| K | 5452 hCoV-19/Australia/VIC14037/2020\|EPI_ISL_563940\|202 | ···················· | ···················· | ···················· | ···················· | NNNNNNNNNNNNNNNNNNNN | ···················· | 1 |
| K | 5453 hCoV-19/Australia/VIC14040/2020\|EPI_ISL_563941\|202 | ···················· | ···················· | ···················· | ···················· | NNNNNNNNNNNNNNNNNNNN | NNNN················ | 1 |
| K | 5455 hCoV-19/Australia/VIC14042/2020\|EPI_ISL_563943\|202 | ···················· | ···················· | ···················· | ···················· | NNNNNNNNNNNNNNNNNNNN | NNNNNNNNNNNNNNNNNNNN | 1 |
| K | 5456 hCoV-19/Australia/VIC14045/2020\|EPI_ISL_563944\|202 | ···················· | ···················· | ···················· | ···················· | NNNNNNNNNNNNNNNNNNNN | NNNNNNNNNNNNNNNNNNNN | 1 |
| K | 5458 hCoV-19/Australia/VIC14048/2020\|EPI_ISL_563946\|202 | ···················· | ···················· | ···················· | ···················· | ···················· | NNNNNNNNN··········· | 1 |
| K | 5461 hCoV-19/Australia/VIC14058/2020\|EPI_ISL_563949\|202 | ···················· | ···················· | ···················· | ···················· | NNNNNNNNNNNNNNNNNNNN | ···················· | 1 |
| K | 5462 hCoV-19/Australia/VIC14060/2020\|EPI_ISL_563950\|202 | ···················· | ···················· | ···················· | ···················· | NNNNNNNNNNNNNNNNNNNN | NNNNNNNNNNNNNNNNNNNN | 1 |
| K | 5467 hCoV-19/Australia/VIC14072/2020\|EPI_ISL_563955\|202 | ···················· | ···················· | ···················· | ···················· | NNNNNNNNNNNNNNNNNNNN | ···················· | 1 |
| K | 5470 hCoV-19/Australia/VIC14079/2020\|EPI_ISL_563958\|202 | ···················· | ···················· | ···················· | ···················· | NNNNNNNNNNNNNNNNNNNN | ···················· | 1 |
| K | 5471 hCoV-19/Australia/VIC14080/2020\|EPI_ISL_563959\|202 | ···················· | ···················· | ···················· | ···················· | NNNNNNNNNNNNNNNNNNNN | NNNNNNNNN··········· | 1 |
| K | 5473 hCoV-19/Australia/VIC14083/2020\|EPI_ISL_563961\|202 | ···················· | ···················· | ···················· | ···················· | NNNNNNNNNNNNNNNNNNNN | NNNNNNNNNNNNNNNNNNNN | 1 |
| K | 5474 hCoV-19/Australia/VIC14085/2020\|EPI_ISL_563962\|202 | ···················· | ···················· | ···················· | ···················· | NNNNNNNNNNNNNNNNNNN· | ···················· | 1 |
| K | 5475 hCoV-19/Australia/VIC14086/2020\|EPI_ISL_563963\|202 | ···················· | ···················· | ···················· | ···················· | NNNN················ | ···················· | 1 |
| K | 5476 hCoV-19/Australia/VIC14087/2020\|EPI_ISL_563964\|202 | ···················· | ···················· | ···················· | ···················· | NNNNNNNNNNNNNNNNNNNN | NNNNNNNNNNNNNNNNN··· | 1 |
| K | 5638 hCoV-19/Gambia/NPHL-760/2020\|EPI_ISL_561327\|2020 | ···················· | ···················· | ···················· | ···················· | NNNNNNNNNNNNNNNNNNNN | NNNNNNNNNNNNNNNNNNNN | 1 |
| K | 5655 hCoV-19/Spain/CN-IBV-97006490/2020\|EPI_ISL_561370\| | ···················· | ···················· | ···················· | ···················· | ···················· | NNNN················ | 1 |
| K | 5657 hCoV-19/Spain/CN-IBV-97006526/2020\|EPI_ISL_561372\| | ···················· | ···················· | ···················· | ···················· | NNNNNNN············· | NNNNNNNNNNNNNNN····· | 1 |
| K | 5660 hCoV-19/Australia/VIC11346/2020\|EPI_ISL_562642\|202 | ···················· | ···················· | ···················· | ···················· | NNNNNNNNNNNNNNNNNNNN | ···················· | 1 |
| K | 5668 hCoV-19/Australia/VIC13719/2020\|EPI_ISL_562650\|202 | ···················· | ···················· | ···················· | ···················· | NNNNNNNNNNNNNNNNNNNN | ···················· | 1 |
| K | 5670 hCoV-19/Australia/VIC13766/2020\|EPI_ISL_562652\|202 | ···················· | ···················· | ···················· | ···················· | NNNNNNNNNNNNNNNNNNNN | ···················· | 1 |
| K | 5672 hCoV-19/Australia/VIC14065/2020\|EPI_ISL_562657\|202 | ···················· | ···················· | ···················· | ···················· | NNNNNNNNNNNNNNNNNNNN | NNNNNNNNNNNNNNNNNNNN | 1 |
| K | 5694 hCoV-19/Australia/VIC9466/2020\|EPI_ISL_562679\|2020 | ···················· | ···················· | ···················· | ···················· | NNNNNNNNNNNNNNNNNNNN | NNNNNNNNNNNNNNNNNNNN | 1 |
| K | 5695 hCoV-19/Australia/VIC9502/2020\|EPI_ISL_562680\|2020 | ···················· | ···················· | ···················· | ···················· | NNNNNNNNNNNNNNNNNNNN | NNNN················ | 1 |
| K | 5706 hCoV-19/Australia/VIC10929/2020\|EPI_ISL_562693\|202 | ···················· | ···················· | ···················· | ···················· | NNNNNNNNNNNNNNNNNNNN | ···················· | 1 |
| K | 5707 hCoV-19/Australia/VIC10938/2020\|EPI_ISL_562694\|202 | ···················· | ···················· | ···················· | ···················· | NNNNNNNNNNNNNNNNNNNN | NNNNNNNNNNNNNNNNNNNN | 1 |
| K | 5709 hCoV-19/Australia/VIC1398/2020\|EPI_ISL_562696\|2020 | ···················· | ···················· | ···················· | ···················· | NNNNNNNNNNNNNNNNNNNN | NNNNNNNNNNNNNNNNNNNN | 1 |
| K | 5710 hCoV-19/Australia/VIC1430/2020\|EPI_ISL_562697\|2020 | ···················· | ···················· | ···················· | ···················· | NNNNNNNNNNNNNNNNNNNN | NNNNNNNNNNNNNNNNNNNN | 1 |
| K | 5714 hCoV-19/Australia/VIC13668/2020\|EPI_ISL_563802\|202 | ···················· | ···················· | ···················· | ···················· | NNNNNNNNNNNNNNNNNNNN | NNNNNNNNN··········· | 1 |
| K | 5718 hCoV-19/Australia/VIC13673/2020\|EPI_ISL_563806\|202 | ···················· | ···················· | ···················· | ···················· | NNNNNNNNNNNNNNNNNNNN | NNNNNNNNNNNNNNNNNNNN | 1 |
| K | 5720 hCoV-19/Australia/VIC1369/2020\|EPI_ISL_563808\|2020 | ···················· | ···················· | ···················· | ···················· | NNNNNNNNNNNNNNNNNNNN | NNNNNNNNNNNNNNNNNNNN | 1 |
| K | 5726 hCoV-19/Australia/VIC13715/2020\|EPI_ISL_563814\|202 | ···················· | ···················· | ···················· | ···················· | NNNNNNNNNNNNNNN····· | ···················· | 1 |
| K | 5731 hCoV-19/Australia/VIC13738/2020\|EPI_ISL_563819\|202 | ···················· | ···················· | ···················· | ···················· | NNNNNNNNNNNNNNNNNNNN | ···················· | 1 |
| K | 5734 hCoV-19/Australia/VIC13744/2020\|EPI_ISL_563822\|202 | ···················· | ···················· | ···················· | ···················· | NNNNNNNNNNNNNNNNNNNN | ···················· | 1 |
| K | 5736 hCoV-19/Australia/VIC13751/2020\|EPI_ISL_563824\|202 | ···················· | ···················· | ···················· | ···················· | NNNNNNNNNNNNNNNNNNNN | ···················· | 1 |
| K | 5742 hCoV-19/Australia/VIC14110/2020\|EPI_ISL_563972\|202 | ···················· | ···················· | ···················· | ···················· | NNNNN··············· | ···················· | 1 |
| K | 5746 hCoV-19/Australia/VIC14117/2020\|EPI_ISL_563976\|202 | ···················· | ···················· | ···················· | ···················· | NNNNNNNNNNNNNNNNNNNN | ···················· | 1 |
| K | 5747 hCoV-19/Australia/VIC1440/2020\|EPI_ISL_563977\|2020 | ···················· | ···················· | ···················· | ···················· | NNNNNNNNNNNNNNNNNNNN | NNNNNNNNNNNNNNNNNNNN | 1 |
| K | 5748 hCoV-19/Australia/VIC2039/2020\|EPI_ISL_563978\|2020 | ···················· | ···················· | ···················· | ···················· | NNNNNNNNNNNNNNNNNNNN | NNNNNNNNNNNNNNNNNNNN | 1 |
| K | 5750 hCoV-19/Australia/VIC2238/2020\|EPI_ISL_563980\|2020 | ···················· | ···················· | ···················· | ···················· | NNNNNNNNNNNNNNNNNNNN | NNNNNNNNNNNNNNNNNNNN | 1 |
| K | 5751 hCoV-19/Australia/VIC2239/2020\|EPI_ISL_563981\|2020 | ···················· | ···················· | ···················· | ···················· | ·NNNNNNNNNNNNNNNNNNN | NNNNNNNNN··········· | 1 |
| K | 5752 hCoV-19/Australia/VIC2240/2020\|EPI_ISL_563982\|2020 | ···················· | ···················· | ···················· | ···················· | NNNNNNNNNNNNNNNNNNNN | ···················· | 1 |
| K | 5753 hCoV-19/Australia/VIC2241/2020\|EPI_ISL_563983\|2020 | ···················· | ···················· | ···················· | ···················· | NNNNNNNNNNNNNNNNNNNN | NNNNNNNNNNNNNNNNNNNN | 1 |
| K | 5754 hCoV-19/Australia/VIC2521/2020\|EPI_ISL_563984\|2020 | ···················· | ···················· | ···················· | ···················· | NNNNNNNNNNNNNNNNNNNN | NNNNNNNNNNNNNNNNNNNN | 1 |
| K | 5755 hCoV-19/Australia/VIC2791/2020\|EPI_ISL_563985\|2020 | ···················· | ···················· | ···················· | ···················· | NNNNNNNNNNNNNNNNNNNN | NNNNNNNNNNNNNNNNNNNN | 1 |
| K | 5756 hCoV-19/Australia/VIC3010/2020\|EPI_ISL_563986\|2020 | ···················· | ···················· | ···················· | ···················· | NNNNNNNNNNNNNNNNNNNN | NNNNNNNNNNNNNNNNNNNN | 1 |

| | | | | | | | | |
|---|---|---|---|---|---|---|---|---|
| K | 5859 hCoV-19/USA/FL-BPHL-1518/2020\|EPI_ISL_568616\|2020- | ···················· | ···················· | ···················· | ···················· | NNNNNNNNNNNNNNNNNNNN | NNNNNNNNNNNNNNNNNNNN | 1 |
| K | 5865 hCoV-19/USA/FL-BPHL-1524/2020\|EPI_ISL_568622\|2020- | ···················· | ···················· | ···················· | ···················· | ·············NNNN··· | ···················· | 1 |
| K | 5929 hCoV-19/Australia/VIC13825/2020\|EPI_ISL_563844\|202 | ···················· | ···················· | ···················· | ···················· | NNNNNNNNNNNNNNNNNNNN | ···················· | 1 |
| K | 5930 hCoV-19/Australia/VIC13826/2020\|EPI_ISL_563845\|202 | ···················· | ···················· | ···················· | ···················· | NNNNNNNNNNNNNNNNNNNN | ···················· | 1 |
| K | 5931 hCoV-19/Australia/VIC13828/2020\|EPI_ISL_563846\|202 | ···················· | ···················· | ···················· | ···················· | NNNNNNNNNNNNNNNN···· | ···················· | 1 |
| K | 6008 hCoV-19/USA/FL-BPHL-1540/2020\|EPI_ISL_568638\|2020- | ···················· | ···················· | ···················· | ···················· | NNNNNNNNNNNNNNNNNNNN | ···················· | 1 |
| K | 6021 hCoV-19/USA/FL-BPHL-1561/2020\|EPI_ISL_568659\|2020- | ···················· | ···················· | ···················· | ···················· | NNNNNNNNNNNNNNNNNNNN | ···················· | 1 |
| K | 6022 hCoV-19/USA/FL-BPHL-1562/2020\|EPI_ISL_568660\|2020- | ···················· | ···················· | ···················· | ···················· | NNNNNNNNNNNNNNNNNNNN | ···········NNNNNNNNN | 1 |
| K | 6025 hCoV-19/USA/FL-BPHL-1565/2020\|EPI_ISL_568663\|2020- | ···················· | ···················· | ···················· | ···················· | NNNNNNNNNNNNNNNNNNNN | ···················· | 1 |
| K | 6033 hCoV-19/USA/FL-BPHL-1577/2020\|EPI_ISL_568675\|2020- | ···················· | ···················· | ···················· | ···················· | NNNNNNNNNNNNNNNNNNNN | ···················· | 1 |
| K | 6084 hCoV-19/USA/WA-UW-12909/2020\|EPI_ISL_570928\|2020-0 | ···················· | ···················· | ···················· | ···················· | ···················· | NNNNNNNNNNNNNNNNNNNN | 1 |
| K | 6209 hCoV-19/Gambia/GC19-3661/2020\|EPI_ISL_561203\|2020- | ···················· | ···················· | ···················· | ···················· | NNNNNNNNNNNNNNNNNNNN | NNNNNNNNNNNNNNNNNNNN | 1 |
| K | 6214 hCoV-19/Gambia/NPHL-1208/2020\|EPI_ISL_561213\|2020- | ···················· | ···················· | ···················· | ···················· | NNNNNNNNNNNNNNNNNNNN | NNNNNNNNNNNNNNNNNNNN | 1 |
| K | 6216 hCoV-19/Gambia/NPHL-1230/2020\|EPI_ISL_561218\|2020- | ···················· | ···················· | ···················· | ···················· | NNNNNNNNNNNNNNNNNNNN | NNNNNNNNNNNNNNNNNNNN | 1 |
| K | 6217 hCoV-19/Gambia/NPHL-1248/2020\|EPI_ISL_561222\|2020- | ···················· | ···················· | ···················· | ···················· | NNNNNNNNNNNNNNNNNNNN | NNNNNNNNNNNNNNNNNNNN | 1 |
| K | 6220 hCoV-19/Gambia/NPHL-1446/2020\|EPI_ISL_561230\|2020- | ···················· | ···················· | ···················· | ···················· | NNNNNNNNNNNNNNNNNNNN | NNNNNNNNNNNNNNNNNNNN | 1 |
| K | 6228 hCoV-19/Gambia/NPHL-2268/2020\|EPI_ISL_561242\|2020 | ···················· | ···················· | ···················· | ···················· | NNNNNNNNNNNNNNNNNNNN | ···················· | 1 |
| K | 6235 hCoV-19/Gambia/NPHL-245/2020\|EPI_ISL_561251\|2020 | ···················· | ···················· | ···················· | ···················· | NNNNNNNNNNNNNNNNNNNN | NNNNNNNNNNNNNNNNNNNN | 1 |
| K | 6258 hCoV-19/Australia/VIC13889/2020\|EPI_ISL_563865\|202 | ···················· | ···················· | ···················· | ···················· | NNNNNNNNNNNN········ | ···················· | 1 |
| K | 6259 hCoV-19/Australia/VIC1389/2020\|EPI_ISL_563866\|2020 | ···················· | ···················· | ···················· | ···················· | NNNNNNNNNNNNNNNNNNNN | NNNNNNNNNNNNNNNNNNNN | 1 |
| K | 6272 hCoV-19/Australia/VIC13910/2020\|EPI_ISL_563879\|202 | ···················· | ···················· | ···················· | ···················· | NNNNNNNNNNNNNNNNNNNN | NNNNNNNNNNNNNNNNNNNN | 1 |
| K | 6278 hCoV-19/Australia/VIC13937/2020\|EPI_ISL_563885\|202 | ···················· | ···················· | ···················· | ···················· | NNNNNNNNNNNNNNNNNNNN | ··K················· | 1 |
| K | 6281 hCoV-19/Australia/VIC13947/2020\|EPI_ISL_563888\|202 | ···················· | ···················· | ···················· | ···················· | NNNNNNNNNNNNNNNN···· | ···················· | 1 |
| K | 6282 hCoV-19/Australia/VIC13948/2020\|EPI_ISL_563889\|202 | ···················· | ···················· | ···················· | ···················· | NNNNNNNNNNNNNNNNNNNN | NNNNNNNNNNNNNNNNNNNN | 1 |
| K | 6284 hCoV-19/Australia/VIC13952/2020\|EPI_ISL_563891\|202 | ···················· | ···················· | ···················· | ···················· | NNNNNNNNNNNNNNNNNNNN | NNNNNNNNNNNNNNNNNNNN | 1 |
| K | 6285 hCoV-19/Australia/VIC13956/2020\|EPI_ISL_563892\|202 | ···················· | ···················· | ···················· | ···················· | NNNNNNNNNNNNNNNNNNNN | NNNNNNNNNNNNNNNNN··· | 1 |
| K | 6287 hCoV-19/Australia/VIC13960/2020\|EPI_ISL_563894\|202 | ···················· | ···················· | ···················· | ···················· | NNNNNNNNNNNNNNNNNNNN | NNN········M········ | 1 |
| K | 6288 hCoV-19/Australia/VIC13962/2020\|EPI_ISL_563895\|202 | ···················· | ···················· | ···················· | ···················· | NNNNNNNNNNNNNNNNNNNN | NNNNNNNNNNNNNNNNNNNN | 1 |
| K | 6289 hCoV-19/Australia/VIC13963/2020\|EPI_ISL_563896\|202 | ···················· | ···················· | ···················· | ···················· | NNNNNNNNNNNNNNNNNNNN | NNNNNNNNNNNNNNNNNNNN | 1 |
| K | 6290 hCoV-19/Australia/VIC13964/2020\|EPI_ISL_563897\|202 | ···················· | ···················· | ···················· | ···················· | NNNNNNNNNNNNNNNNNNNN | NNNNNNNNNNNNNNNN···· | 1 |
| K | 6291 hCoV-19/Australia/VIC13965/2020\|EPI_ISL_563898\|202 | ···················· | ···················· | ···················· | ···················· | ···················· | NNNN················ | 1 |
| K | 6292 hCoV-19/Australia/VIC13966/2020\|EPI_ISL_563899\|202 | ···················· | ···················· | ···················· | ···················· | NNNNNNNNNNNNNNNNNNNN | NNNNNNNNNNNNNNNNNNNN | 1 |
| K | 6325 hCoV-19/Australia/VIC6947/2020\|EPI_ISL_564160\|2020 | ···················· | ···················· | ···················· | ···················· | NNNNNNNNNNNNNNNNNNNN | NNNNNNNNNNNNNNNN···· | 1 |
| K | 6326 hCoV-19/Australia/VIC6950/2020\|EPI_ISL_564161\|2020 | ···················· | ···················· | ···················· | ···················· | NNNNNNNNNNNNNNNNNNNN | ···················· | 1 |
| K | 6329 hCoV-19/Australia/VIC6964/2020\|EPI_ISL_564164\|2020 | ···················· | ···················· | ···················· | ···················· | ···················· | NNNN················ | 1 |
| K | 6416 hCoV-19/Australia/VIC6982/2020\|EPI_ISL_564178\|2020 | ···················· | ···················· | ···················· | ···················· | NNNNNNNNNNNNNNNNNNNN | ···················· | 1 |
| K | 6417 hCoV-19/Australia/VIC6983/2020\|EPI_ISL_564179\|2020 | ···················· | ···················· | ···················· | ···················· | NNNNNNNNNNNNNNNNNNNN | NNNNNNNNNNNNNNNNNNNN | 1 |
| K | 6418 hCoV-19/Australia/VIC6984/2020\|EPI_ISL_564180\|2020 | ···················· | ···················· | ···················· | ···················· | NNNNNNNNNNNNNNNNNNNN | NNNNNNNNN·····NNNNNN | 1 |
| K | 6419 hCoV-19/Australia/VIC6985/2020\|EPI_ISL_564181\|2020 | ···················· | ···················· | ···················· | ···················· | NNNNNNNNNNNNNNNNN··· | NNNNNNNNNNNNNNNNNNNN | 1 |
| K | 6423 hCoV-19/Australia/VIC6991/2020\|EPI_ISL_564185\|2020 | ···················· | ···················· | ···················· | ···················· | NNNNNNNNNNNNNN······ | NNNN·NNNN··········· | 1 |
| K | 6424 hCoV-19/Australia/VIC6992/2020\|EPI_ISL_564186\|2020 | ···················· | ···················· | ···················· | ···················· | NNNNNNNNNNNNNNNNNNNN | ···················· | 1 |
| K | 6425 hCoV-19/Australia/VIC6993/2020\|EPI_ISL_564187\|2020 | ···················· | ···················· | ···················· | ···················· | NNNNNNNNNNNNNNNNNNNN | NNNNNNNNNNNNNNNNNNNN | 1 |
| K | 6430 hCoV-19/Australia/VIC7000/2020\|EPI_ISL_564192\|2020 | ···················· | ···················· | ···················· | ···················· | NNNNNNNNNNNNNNNNNNNN | ···················· | 1 |
| K | 6431 hCoV-19/Australia/VIC7001/2020\|EPI_ISL_564193\|2020 | ···················· | ···················· | ···················· | ···················· | NNNNNNNNNNNNNNNNNNNN | NNNNN··············· | 1 |
| K | 6434 hCoV-19/Australia/VIC7008/2020\|EPI_ISL_564196\|2020 | ···················· | ···················· | ···················· | ···················· | NNNNNNNNNNNN········ | NNNNNNNNNNNNNNNN···· | 1 |
| K | 6436 hCoV-19/Australia/VIC7018/2020\|EPI_ISL_564198\|2020 | ···················· | ···················· | ···················· | ···················· | NNNNNNNNNNNNNNNNNNNN | ···················· | 1 |
| K | 6437 hCoV-19/Australia/VIC7026/2020\|EPI_ISL_564199\|2020 | ···················· | ···················· | ···················· | ···················· | NNNNNNNNNNNNNNNNNNNN | NNNNNNNNNNNNNNNNNNNN | 1 |
| K | 6441 hCoV-19/Australia/VIC7036/2020\|EPI_ISL_564203\|2020 | ···················· | ···················· | ···················· | ···················· | NNNNNNNNNNNNNNNNNNNN | NNNNN··············· | 1 |
| K | 6442 hCoV-19/Australia/VIC7040/2020\|EPI_ISL_564204\|2020 | ···················· | ···················· | ···················· | ···················· | NNNNNNNNNNNNNNNNNNNN | ··················NN | 1 |
| K | 6446 hCoV-19/Australia/VIC7047/2020\|EPI_ISL_564208\|2020 | ···················· | ···················· | ···················· | ···················· | NNNNNNNNNNNNNNNNNNNN | NNNNNNNNNNNNNNNNNNNN | 1 |
| K | 6449 hCoV-19/Australia/VIC7054/2020\|EPI_ISL_564211\|2020 | ···················· | ···················· | ···················· | ···················· | NNNNNNNNNNNNNNNNNNNN | ···················· | 1 |
| K | 6453 hCoV-19/Australia/VIC7072/2020\|EPI_ISL_564215\|2020 | ···················· | ···················· | ···················· | ···················· | NNNNNNNNNNNNNNNNN··· | ···················· | 1 |
| K | 6454 hCoV-19/Australia/VIC7073/2020\|EPI_ISL_564216\|2020 | ···················· | ···················· | ···················· | ···················· | NNNNNNNNNNNNNNNNNNNN | ···················· | 1 |
| K | 6546 hCoV-19/Australia/VIC9454/2020\|EPI_ISL_565558\|2020 | ···················· | ···················· | ···················· | ···················· | NNNNNNNNNNNNNNNNNNNN | NNNNNNNNNNNNNNNNNNNN | 1 |
| K | 6551 hCoV-19/Australia/VIC9467/2020\|EPI_ISL_565563\|2020 | ···················· | ···················· | ···················· | ···················· | ··NNNNNNNNNNNNNNNNNN | ···················· | 1 |
| K | 6552 hCoV-19/Australia/VIC9474/2020\|EPI_ISL_565564\|2020 | ···················· | ···················· | ···················· | ···················· | NNNNNNNNNNNNNNNNNNNN | NNNN··NNNNNNNNNNNNNN | 1 |
| K | 6561 hCoV-19/Australia/VIC9492/2020\|EPI_ISL_565573\|2020 | ···················· | ···················· | ···················· | ···················· | NNNNNNNNNNNNNNNNNNNN | NNNNNNNNNNNNNNNNNNNN | 1 |
| K | 6595 hCoV-19/England/MILK-9C30B5/2020\|EPI_ISL_566853\|20 | ···················· | ···················· | ···················· | NNNNNNNNNNNNNNNNNNNN | ···················· | ···················· | 1 |
| K | 6681 hCoV-19/Switzerland/VD-ETHZ-280207/2020\|EPI_ISL_56 | ···················· | ···················· | ···················· | ···················· | ·········K·MK···M··M | ···················· | 1 |
| K | 6718 hCoV-19/Australia/VIC6715/2020\|EPI_ISL_564006\|2020 | ···················· | ···················· | ···················· | ···················· | NNNNNNNNNNNNNNNNNNNN | NNNNNNNNNNNNNNNNNNNN | 1 |
| K | 6749 hCoV-19/Australia/VIC9010/2020\|EPI_ISL_565353\|2020 | ···················· | ···················· | ···················· | ···················· | NNNNNNNNNNNNNNNNNNNN | ···················· | 1 |
| K | 6931 hCoV-19/Australia/VIC6761/2020\|EPI_ISL_564037\|2020 | ···················· | ···················· | ···················· | ···················· | NNNN················ | NNNN················ | 1 |
| K | 6932 hCoV-19/Australia/VIC6762/2020\|EPI_ISL_564038\|2020 | ···················· | ···················· | ···················· | ········S··········· | NNNNNNNNNNNNNNNNNNNN | ···················· | 1 |
| K | 6941 hCoV-19/Australia/VIC6774/2020\|EPI_ISL_564047\|2020 | ···················· | ···················· | ···················· | ···················· | NNNNNNNNNNNNNNNNNNNN | ···················· | 1 |
| K | 6944 hCoV-19/Australia/VIC6779/2020\|EPI_ISL_564050\|2020 | ···················· | ···················· | ···················· | ···················· | ···················· | NNNNNNNNNNNNNNNNNNNN | 1 |
| K | 6945 hCoV-19/Australia/VIC6781/2020\|EPI_ISL_564051\|2020 | ···················· | ···················· | ···················· | ···················· | ·NNNNNNNNNNNNNNNNNNN | NNNNNNNNNNNNNNNN···· | 1 |
| K | 6970 hCoV-19/Australia/VIC6818/2020\|EPI_ISL_564076\|2020 | ···················· | ···················· | ···················· | ···················· | NNNNNNNNNNNNNNNNNNNN | ···················· | 1 |
| K | 6971 hCoV-19/Australia/VIC6824/2020\|EPI_ISL_564077\|2020 | ···················· | ···················· | ···················· | ···················· | NNNNNNNNNNNNNNNNNNNN | NNNNNNNNN···NNN··NNN | 1 |
| K | 6980 hCoV-19/Australia/VIC6834/2020\|EPI_ISL_564086\|2020 | ···················· | ···················· | ···················· | ···················· | NNNNNNNNNNNNNNNNNNNN | NNNNNNNNNNNNNNNNNNNN | 1 |
| K | 6999 hCoV-19/Australia/VIC8992/2020\|EPI_ISL_565342\|2020 | ···················· | ·····A·············· | ···················· | ···················· | NNNNNNNNNNNNNNNNNNNN | ···R················ | 1 |
| K | 7005 hCoV-19/Australia/VIC9023/2020\|EPI_ISL_565359\|2020 | ···················· | ···················· | ···················· | ···················· | ···················· | NNNNNNNNNNNNNNNN···· | 1 |
| K | 7256 hCoV-19/Australia/VIC6900/2020\|EPI_ISL_564125\|2020 | ···················· | ···················· | ···················· | ···················· | NNNNNNNNNNNNNNNNNNNN | ···················· | 1 |
| K | 7486 hCoV-19/Australia/VIC8927/2020\|EPI_ISL_565284\|2020 | ···················· | ···················· | ···················· | ···················· | NNNNNNNNNNNNNNNNNNNN | ···················· | 1 |
| K | 7500 hCoV-19/Australia/VIC8944/2020\|EPI_ISL_565298\|2020 | ···················· | ···················· | ···················· | ···················· | ···················· | NNNNNNNNN··········· | 1 |
| K | 7504 hCoV-19/Australia/VIC8949/2020\|EPI_ISL_565303\|2020 | ···················· | ···················· | ···················· | ···················· | NNNNNNNNNNNNNNNNNNNN | ···················· | 1 |
| K | 7525 hCoV-19/Australia/VIC8973/2020\|EPI_ISL_565324\|2020 | ···················· | ···················· | ···················· | ···················· | NNNNNNNNNNNNNNNNNNNN | ···················· | 1 |
| K | 7532 hCoV-19/Australia/VIC8981/2020\|EPI_ISL_565331\|2020 | ···················· | ···················· | ···················· | ···················· | NNNNNNNNNNNN········ | ···················· | 1 |
| K | 7535 hCoV-19/Australia/VIC8984/2020\|EPI_ISL_565334\|2020 | ···················· | ·····R·············· | ···················· | ···················· | NNNNNNNNNNNNNNNNNNNN | NNNNNNNNNNNNNNNNNNNN | 1 |
| K | 7686 hCoV-19/Australia/VIC10413/2020\|EPI_ISL_562905\|202 | ···················· | ···················· | ···················· | ···················· | NNNNNNN············· | ···················· | 1 |
| K | 7687 hCoV-19/Australia/VIC10419/2020\|EPI_ISL_562906\|202 | ···················· | ···················· | ···················· | ···················· | NNNNNNNNNNNNNNNNNNNN | ···················· | 1 |

| | | | | | | | | | |
|---|---|---|---|---|---|---|---|---|---|
| K | 7688 | hCoV-19/Australia/VIC10421/2020\|EPI_ISL_562907\|202 | ···················· | ···················· | ···················· | ···················· | NNNNNNNNNNNNNNNNNNNN | NNNNNNNNNNNNNNNNNNNN | 1 |
| K | 7689 | hCoV-19/Australia/VIC10422/2020\|EPI_ISL_562908\|202 | ···················· | ···················· | ···················· | ···················· | NNNNNNNNNNNNNNNNNNNN | NNNNNNNNNNNNNNNNNNNN | 1 |
| K | 7690 | hCoV-19/Australia/VIC10426/2020\|EPI_ISL_562909\|202 | ···················· | ···················· | ···················· | ···················· | NNNNNNNNNNNNNNNNNNNN | ···················· | 1 |
| K | 7699 | hCoV-19/Australia/VIC10457/2020\|EPI_ISL_562918\|202 | ···················· | ···················· | ···················· | ···················· | ···················· | NNNNN··············· | 1 |
| K | 7702 | hCoV-19/Australia/VIC10464/2020\|EPI_ISL_562921\|202 | ···················· | ···················· | ···················· | ···················· | NNNNNNNNNNNN········ | NNNN················ | 1 |
| K | 7707 | hCoV-19/Australia/VIC10472/2020\|EPI_ISL_562926\|202 | ···················· | ···················· | ···················· | ···················· | NNNNNNNNNNNNNNNNNNNN | NNNNNNNNNNNNNNNNNNNN | 1 |
| K | 7718 | hCoV-19/Australia/VIC10504/2020\|EPI_ISL_562937\|202 | ···················· | ···················· | ···················· | ···················· | NNNNNNNNNNNNNNNNN··· | ···················· | 1 |
| K | 7725 | hCoV-19/Australia/VIC10521/2020\|EPI_ISL_562944\|202 | ···················· | ···················· | ···················· | ···················· | NNNNNNNNNNNNNNNNNNNN | ···················· | 1 |
| K | 7747 | hCoV-19/Australia/VIC10576/2020\|EPI_ISL_562966\|202 | ···················· | ···················· | ···················· | ···················· | NNNNNNNNNNNNNNNNNNNN | NNNNNNNNNNNNNNNNNNNN | 1 |
| K | 7754 | hCoV-19/Australia/VIC10589/2020\|EPI_ISL_562973\|202 | ···················· | ···················· | ···················· | ···················· | NNNNNNNNNNNNNNNNNNNN | ···················· | 1 |
| K | 7757 | hCoV-19/Australia/VIC10594/2020\|EPI_ISL_562976\|202 | ···················· | ···················· | ···················· | ···················· | NNNNNNNNNNNNNNNNNNNN | ···················· | 1 |
| K | 7759 | hCoV-19/Australia/VIC10597/2020\|EPI_ISL_562978\|202 | ···················· | ···················· | ···················· | ···················· | NNNNNNNNNNNNNNNNNNNN | ···················· | 1 |
| K | 7762 | hCoV-19/Australia/VIC10600/2020\|EPI_ISL_562981\|202 | ···················· | ···················· | ···················· | ···················· | ···················· | NNNNNNNNNNNNNNNNNNNN | 1 |
| K | 7818 | hCoV-19/Australia/VIC8719/2020\|EPI_ISL_565155\|2020 | ·······Y············ | ···················· | ···················· | ····NN·············· | NNNNNNNNNNNNNNNNNNNN | NNNNNNNNNNNNNNNNNNNN | 1 |
| K | 7823 | hCoV-19/Australia/VIC8759/2020\|EPI_ISL_565180\|2020 | ···················· | ···················· | ···················· | ···················· | NNNNNNNNNNNNNNNNNNNN | ···················· | 1 |
| K | 7831 | hCoV-19/Australia/VIC8777/2020\|EPI_ISL_565188\|2020 | ···················· | ···················· | ···················· | ···················· | NNNNNNNNNNNNNNNNNNNN | ···················· | 1 |
| K | 7838 | hCoV-19/Australia/VIC8787/2020\|EPI_ISL_565195\|2020 | ···················· | ···················· | ···················· | ···················· | NNNNNNNNNNNNNNNNNNNN | NNNNNNNNNNNNNNNNNNNN | 1 |
| K | 7961 | hCoV-19/Australia/VIC10604/2020\|EPI_ISL_562984\|202 | ···················· | ···················· | ···················· | ···················· | NNNNNNNNNNNN········ | ···················· | 1 |
| K | 7963 | hCoV-19/Australia/VIC10608/2020\|EPI_ISL_562986\|202 | ···················· | ···················· | ···················· | ···················· | ·NNNNNNNNNN········· | ···················· | 1 |
| K | 7967 | hCoV-19/Australia/VIC10612/2020\|EPI_ISL_562990\|202 | ···················· | ···················· | ···················· | ···················· | NNNNNNNNNNNNNNNNNNNN | NNNNNNNNNNNNNNNNNNNN | 1 |
| K | 7968 | hCoV-19/Australia/VIC10613/2020\|EPI_ISL_562991\|202 | ···················· | ···················· | ···················· | ···················· | ···················· | NNNN················ | 1 |
| K | 7972 | hCoV-19/Australia/VIC10620/2020\|EPI_ISL_562995\|202 | ···················· | ···················· | ···················· | ···················· | NNNNNNNNNNNNNNNNNNNN | NNNNNNNNNNNNNNNNNNNN | 1 |
| K | 7973 | hCoV-19/Australia/VIC10621/2020\|EPI_ISL_562996\|202 | ···················· | ···················· | ···················· | ···················· | NNNNNNNNNN·········· | ···················· | 1 |
| K | 7975 | hCoV-19/Australia/VIC10623/2020\|EPI_ISL_562998\|202 | ···················· | ···················· | ···················· | ···················· | NNNNNNNNNNNN········ | ···················· | 1 |
| K | 7976 | hCoV-19/Australia/VIC10624/2020\|EPI_ISL_562999\|202 | ···················· | ···················· | ···················· | ···················· | NNNNNNNNNNNNNNNNNNNN | ···················· | 1 |
| K | 8003 | hCoV-19/Australia/VIC8804/2020\|EPI_ISL_565206\|2020 | ···················· | ···················· | ···················· | ···················· | ·············NNNNNNN | ···················· | 1 |
| L | 259 | hCoV-19/Wales/PHWC-16C001/2020\|EPI_ISL_574002\|2020 | ···················· | ···················· | ···················· | ···················· | ···················· | NNNNNNNNNNNNNNN····· | 1 |
| L | 263 | hCoV-19/Wales/PHWC-16C05C/2020\|EPI_ISL_574006\|2020 | ···················· | ···················· | ···················· | ···················· | NNNNNNNNNNNN········ | ···················· | 1 |
| L | 280 | hCoV-19/Wales/PHWC-16C1E0/2020\|EPI_ISL_574023\|2020 | ···················· | ···················· | ···················· | ···················· | NNNN················ | ···················· | 1 |
| L | 289 | hCoV-19/Wales/PHWC-16C292/2020\|EPI_ISL_574032\|2020 | ···················· | ···················· | ···················· | ···················· | NNNN················ | ···················· | 1 |
| L | 292 | hCoV-19/Wales/PHWC-16C2CF/2020\|EPI_ISL_574035\|2020 | ···················· | ···················· | ···················· | ···················· | NNNNNNNNNNNNNNNNNNNN | NNNNNNNNNNNNNNNNNNNN | 1 |
| L | 296 | hCoV-19/Wales/PHWC-16C308/2020\|EPI_ISL_574039\|2020 | ···················· | ···················· | ···················· | ···················· | NNNNNN·············· | ···················· | 1 |
| L | 587 | hCoV-19/England/ALDP-9CC1C8/2020\|EPI_ISL_572940\|20 | ···················· | ···················· | ···················· | ···················· | NNNNNNNNNNNNNNNNNNNN | NNNNNNNNNNNNNNNNNNNN | 1 |
| L | 589 | hCoV-19/England/MILK-9C700C/2020\|EPI_ISL_572942\|20 | ···················· | ···················· | ···················· | ···················· | NNNNNNNNNNNNNNNNNNNN | NNNNNNNNNNNNNNNNNNNN | 1 |
| L | 599 | hCoV-19/England/ALDP-9CC59F/2020\|EPI_ISL_572954\|20 | ···················· | ···················· | ···················· | ···················· | NNNNNNNNNNNNNNNNNNNN | NNNNNNNNNNNNNNNNNNNN | 1 |
| L | 788 | hCoV-19/England/ALDP-9CC0F8/2020\|EPI_ISL_572939\|20 | ···················· | ·---················ | ···················· | ···················· | NNNNNNNNNNNNNNNNNNNN | NNNNNNNNNNNNNNNNNNNN | 1 |
| L | 1103 | hCoV-19/England/ALDP-9CC535/2020\|EPI_ISL_572847\|20 | ···················· | ···················· | ···················· | ···················· | NNNNNNNNNNNNNNNNNNNN | NNNNNNNNNNNNNNNNNNNN | 1 |
| L | 1110 | hCoV-19/Wales/PHWC-16C97F/2020\|EPI_ISL_572854\|2020 | ···················· | ···················· | ···················· | ···················· | ·NNNNNNNNNNNNNNNNNNN | ···················· | 1 |
| L | 1118 | hCoV-19/England/CAMB-1B5F75/2020\|EPI_ISL_572862\|20 | ···················· | ···················· | ···················· | ···················· | NNNNNNNNNNNNNNNNNNNN | NNNNNNNNNNNNNNNNNNNN | 1 |
| L | 1137 | hCoV-19/Wales/PHWC-16C511/2020\|EPI_ISL_572881\|2020 | ···················· | ···················· | ···················· | ···················· | NNNNNNNNNNNNNN······ | ···················· | 1 |
| L | 1144 | hCoV-19/Wales/PHWC-16D0C4/2020\|EPI_ISL_572888\|2020 | ···················· | ···················· | ···················· | ···················· | NNNNNNNNNNNNNNNNNNNN | ···················· | 1 |
| L | 1594 | hCoV-19/England/ALDP-9CC614/2020\|EPI_ISL_572711\|20 | ···················· | ···················· | ···················· | ···················· | NNNNNNNNNNNNNNNNNNNN | NNNNNNNNNNNNNNNNNNNN | 1 |
| L | 1600 | hCoV-19/England/NORW-EF16E/2020\|EPI_ISL_572717\|202 | ···················· | ···················· | ···················· | ···················· | NNNNNNNNNNNNNNNNNNNN | NNNNNNNNNNNNNNNNNNNN | 1 |
| L | 1601 | hCoV-19/England/NORW-EF632/2020\|EPI_ISL_572718\|202 | ···················· | ···················· | ···················· | ···················· | NNNNNNNNNNNNNNNNNNNN | NNNNNNNNNNNNNNNNNNNN | 1 |
| L | 1603 | hCoV-19/England/NORW-EF82D/2020\|EPI_ISL_572720\|202 | ···················· | ···················· | ···················· | ···················· | NNNNNNNNNNNNNNNNNNNN | NNNNNNNNNNNNNNNNNNNN | 1 |
| L | 1766 | hCoV-19/USA/MI-UM-10035929654/2020\|EPI_ISL_583400\| | ···················· | ···················· | ···················· | ···················· | NNNNNNNNNNNNNNNNNNNN | ···················· | 1 |
| L | 1768 | hCoV-19/USA/MI-UM-10035923993/2020\|EPI_ISL_583402\| | ···················· | ·············A······ | ···················· | ···················· | NNNNNNNNNNNNNNNNNNNN | NNNNNNNNNNNNNNNNNNNN | 1 |
| L | 1769 | hCoV-19/USA/MI-UM-10036114947/2020\|EPI_ISL_583404\| | T··················· | ·A·················· | ···················· | ···················· | NNNNNNNNNNNNNNNNNNNN | ···················· | 1 |
| L | 1770 | hCoV-19/USA/MI-UM-10036110847/2020\|EPI_ISL_583405\| | ···················· | ···················· | ···················· | ···················· | NNNNNNNNNNNNNNNNNNNN | ···················· | 1 |
| L | 1771 | hCoV-19/USA/MI-UM-10036102177/2020\|EPI_ISL_583406\| | ···················· | ·A·················· | ···················· | ···················· | NNNNNNNNNNNNNNNNNNNN | ···················· | 1 |
| L | 1774 | hCoV-19/USA/MI-UM-10036102583/2020\|EPI_ISL_583409\| | T··················· | ·A·················· | ···················· | ···················· | NNNNNNNNNNNNNNNNNNNN | ···················· | 1 |
| L | 1775 | hCoV-19/USA/MI-UM-10036061167/2020\|EPI_ISL_583411\| | ···················· | ···················· | ···················· | ···················· | NNNNNNNNNNNNNNNNNNNN | NNNNNNNNNNNNNNNNNNNN | 1 |
| L | 1776 | hCoV-19/USA/MI-UM-10036095408/2020\|EPI_ISL_583412\| | T··················· | ·A·················· | ···················· | ···················· | NNNNNNNNNNNNNNNNNNNN | NNNNNNNNNNNNNNNNNNNN | 1 |
| L | 1778 | hCoV-19/USA/MI-UM-10036055944/2020\|EPI_ISL_583414\| | ···················· | ···················· | ···················· | ···················· | NNNNNNNNNNNNNNNNNNNN | NNNNNNNNNNNNNNNNNNNN | 1 |
| L | 1779 | hCoV-19/USA/MI-UM-10036095605/2020\|EPI_ISL_583415\| | T··················· | ·A·················· | ···················· | ···················· | NNNNNNNNNNNNNNNNNNNN | ···················· | 1 |
| L | 1780 | hCoV-19/USA/MI-UM-10036077350/2020\|EPI_ISL_583416\| | ···················· | ···················· | ···················· | ···················· | NNNNNNNNNNNNNNNNNNNN | ···················· | 1 |
| L | 1781 | hCoV-19/USA/MI-UM-10036058805/2020\|EPI_ISL_583417\| | ···················· | ···················· | ···················· | ···················· | NNNNNNNNNNNNNNNNNNNN | ···················· | 1 |
| L | 1782 | hCoV-19/USA/MI-UM-10036075143/2020\|EPI_ISL_583418\| | ···················· | ···················· | ···················· | ···················· | NNNNNNNNNNNNNNNNNNNN | NNNNNNNNNNNNNNNNNNNN | 1 |
| L | 1784 | hCoV-19/USA/MI-UM-10036116103/2020\|EPI_ISL_583420\| | ···················· | ···················· | ···················· | ···················· | NNNNNNNNNNNNNNNNNNNN | NNNNNNNNNNNNNNNNNNNN | 1 |
| L | 1785 | hCoV-19/USA/MI-UM-10036138856/2020\|EPI_ISL_583422\| | ···················· | ···················· | ···················· | ···················· | NNNNNNNNNNNNNNNNNNNN | NNNNNNNNNNNNNNNNNNNN | 1 |
| L | 1786 | hCoV-19/USA/MI-UM-10036138458/2020\|EPI_ISL_583423\| | ···················· | ···················· | ···················· | ···················· | NNNNNNNNNNNNNNNNNNNN | ···················· | 1 |
| L | 1788 | hCoV-19/USA/MI-UM-10036130662/2020\|EPI_ISL_583425\| | ···················· | ·············A······ | ···················· | ···················· | NNNNNNNNNNNNNNNNNNNN | ···················· | 1 |
| L | 1893 | hCoV-19/India/MH-NIV-214342/2020\|EPI_ISL_577676\|20 | ···················· | ···················· | ···················· | NNNNNNNNNNNNNN······ | ···················· | ···················· | 1 |
| L | 1981 | hCoV-19/Spain/CB-IBV-98007074/2020\|EPI_ISL_582104\| | ···················· | ···················· | ···················· | ···················· | NNNNNNNNNNNNNNNNNNNN | ···················· | 1 |
| L | 2051 | hCoV-19/England/ALDP-9CC0E9/2020\|EPI_ISL_573085\|20 | ···················· | ···················· | ···················· | ···················· | NNNNNNNNNNNNNNNNNNNN | NNNNNNNNNNNNNNNNNNNN | 1 |
| L | 2052 | hCoV-19/England/ALDP-9CC104/2020\|EPI_ISL_573086\|20 | ···················· | ···················· | ···················· | ···················· | NNNNNNNNNNNNNNNNNNNN | NNNNNNNNNNNNNNNNNNNN | 1 |
| L | 2053 | hCoV-19/England/ALDP-9CC122/2020\|EPI_ISL_573087\|20 | ···················· | ···················· | ···················· | ···················· | ···················· | NNNNNNNNN··········· | 1 |
| L | 2055 | hCoV-19/England/ALDP-9CC1B9/2020\|EPI_ISL_573090\|20 | ···················· | ···················· | ···················· | ···················· | NNNNNNNNNNNNNNNNNNNN | NNNNNNNNNNNNNNNNNNNN | 1 |
| L | 2056 | hCoV-19/England/ALDP-9CC1F5/2020\|EPI_ISL_573091\|20 | ···················· | ···················· | ···················· | ···················· | ···················· | NNNNNNNNNNNNNNNNNNNN | 1 |
| L | 2058 | hCoV-19/England/ALDP-9CC26B/2020\|EPI_ISL_573093\|20 | ···················· | ···················· | ···················· | ···················· | NNNNNNNNNNNNNNNNNNNN | NNNNNNNNNNNNNNNNNNNN | 1 |
| L | 2059 | hCoV-19/England/ALDP-9CC289/2020\|EPI_ISL_573094\|20 | ···················· | ···················· | ···················· | ···················· | NNNNNNNNNNNNNNNNNNNN | NNNNNNNNNNNNNNNNNNNN | 1 |
| L | 2060 | hCoV-19/England/ALDP-9CC2B6/2020\|EPI_ISL_573095\|20 | ···················· | ···················· | ···················· | ···················· | NNNNNNNNNNNNNNNNNNNN | NNNNNNNNNNNNNNNNNNNN | 1 |
| L | 2062 | hCoV-19/England/ALDP-9CC30E/2020\|EPI_ISL_573097\|20 | ···················· | ···················· | ···················· | ···················· | ······NNNNNNNNNNNNNN | NNNNNNNNNNNNNNNNNNNN | 1 |
| L | 2064 | hCoV-19/England/ALDP-9CC32C/2020\|EPI_ISL_573099\|20 | ···················· | ···················· | ···················· | ···················· | NNNNNNNNNNNNNNNNNNNN | NNNNNNNNNNNNNNNNNNNN | 1 |
| L | 2074 | hCoV-19/New Zealand/20VR2060/2020\|EPI_ISL_579220\|2 | ···················· | ···················· | ···················· | ···················· | NNNNNNNNNNNNNNNNNNNN | NNNNNNNNNNNNNNNNNNNN | 1 |
| L | 2075 | hCoV-19/New Zealand/20VR2069/2020\|EPI_ISL_579223\|2 | ···················· | ···················· | ···················· | ···················· | NNNNNNNNNNNNNNNNNNNN | NNNNNNNNNNNNNNNNNNNN | 1 |
| L | 2076 | hCoV-19/New Zealand/20VR2080/2020\|EPI_ISL_579225\|2 | ···················· | ···················· | ···················· | ···················· | NNNNNNNNNNNNNNNNNNNN | NNNNNNNNNNNNNNNNNNNN | 1 |
| L | 2077 | hCoV-19/New Zealand/20VR2082/2020\|EPI_ISL_579226\|2 | ···················· | ···················· | ···················· | ···················· | NNNNNNNNNNNNNNNNNNNN | NNNNNNNNNNNNNNNNNNNN | 1 |
| L | 2079 | hCoV-19/New Zealand/20VR2100/2020\|EPI_ISL_579228\|2 | ···················· | ···················· | ···················· | ···················· | NNNNNNNNNNNNNNNNNNNN | NNNNNNNNNNNNNNNNNNNN | 1 |
| L | 2085 | hCoV-19/New Zealand/20VR2478/2020\|EPI_ISL_579239\|2 | ···················· | ···················· | ···················· | ···················· | NNNNNNNNNNNNNNNNNNNN | NNNNNNNNNNNNNNNNNNNN | 1 |

| | | | | | | | | | |
|---|---|---|---|---|---|---|---|---|---|
| L | 2096 | hCoV-19/New Zealand/20VR2499/2020\|EPI_ISL_579254\|2 | ···················· | ···················· | ···················· | ···················· | NNNNNNNNNNNNNNNNNNNN | NNNNNNNNNNNNNNNNNNNN | 1 |
| L | 2098 | hCoV-19/New Zealand/20VR2506/2020\|EPI_ISL_579260\|2 | ···················· | ···················· | ···················· | ···················· | NNNNNNNNNNNNNNNNNNNN | NNNNNNNNNNNNNNNNNNNN | 1 |
| L | 2100 | hCoV-19/New Zealand/20VR2516/2020\|EPI_ISL_579262\|2 | ···················· | ···················· | ···················· | ···················· | NNNNNNNNNNNNNNNNNNNN | NNNNNNNNNNNNNNNNNNNN | 1 |
| L | 2102 | hCoV-19/New Zealand/20VR2518/2020\|EPI_ISL_579264\|2 | ···················· | ···················· | ···················· | ···················· | NNNNNNNNNNNNNNNNNNNN | NNNNNNNNNNNNNNNNNNNN | 1 |
| L | 2105 | hCoV-19/New Zealand/20VR2521/2020\|EPI_ISL_579267\|2 | ···················· | ···················· | ···················· | ···················· | NNNNNNNNNNNNNNNNNNNN | NNNNNNNNNNNNNNNNNNNN | 1 |
| L | 2106 | hCoV-19/New Zealand/20VR2523/2020\|EPI_ISL_579268\|2 | ···················· | ···················· | ···················· | ···················· | NNNNNNNNNNNNNNNNNNNN | NNNNNNNNNNNNNNNNNNNN | 1 |
| L | 2107 | hCoV-19/New Zealand/20VR2524/2020\|EPI_ISL_579269\|2 | ···················· | ···················· | ···················· | ···················· | NNNNNNNNNNNNNNNNNNNN | NNNNNNNNNNNNNNNNNNNN | 1 |
| L | 2108 | hCoV-19/New Zealand/20VR2525/2020\|EPI_ISL_579270\|2 | ···················· | ···················· | ···················· | ···················· | NNNNNNNNNNNNNNNNNNNN | NNNNNNNNNNNNNNNNNNNN | 1 |
| L | 2111 | hCoV-19/New Zealand/20VR2528/2020\|EPI_ISL_579273\|2 | ···················· | ···················· | ···················· | ···················· | NNNNNNNNNNNNNNNNNNNN | NNNNNNNNNNNNNNNNNNNN | 1 |
| L | 2112 | hCoV-19/New Zealand/20VR2529/2020\|EPI_ISL_579274\|2 | ···················· | ···················· | ···················· | ···················· | NNNNNNNNNNNNNNNNNNNN | NNNNNNNNNNNNNNNNNNNN | 1 |
| L | 2115 | hCoV-19/New Zealand/20VR2534/2020\|EPI_ISL_579279\|2 | ···················· | ···················· | ···················· | ···················· | NNNNNNNNNNNNNNNNNNNN | NNNNNNNNNNNNNNNNNNNN | 1 |
| L | 2117 | hCoV-19/New Zealand/20VR2536/2020\|EPI_ISL_579281\|2 | ···················· | ···················· | ···················· | ···················· | NNNNNNNNNNNNNNNNNNNN | NNNNNNNNNNNNNNNNNNNN | 1 |
| L | 2118 | hCoV-19/New Zealand/20VR2537/2020\|EPI_ISL_579282\|2 | ···················· | ···················· | ···················· | ···················· | NNNNNNNNNNNNNNNNNNNN | NNNNNNNNNNNNNNNNNNNN | 1 |
| L | 2120 | hCoV-19/New Zealand/20VR2540/2020\|EPI_ISL_579284\|2 | ···················· | ···················· | ···················· | ···················· | NNNNNNNNNNNNNNNNNNNN | NNNNNNNNNNNNNNNNNNNN | 1 |
| L | 2121 | hCoV-19/New Zealand/20VR2543/2020\|EPI_ISL_579286\|2 | ···················· | ···················· | ···················· | ···················· | NNNNNNNNNNNNNNNNNNNN | NNNNNNNNNNNNNNNNNNNN | 1 |
| L | 2131 | hCoV-19/New Zealand/20VR2559/2020\|EPI_ISL_579298\|2 | ···················· | ···················· | ···················· | ···················· | NNNNNNNNNNNNNNNNNNNN | NNNNNNNNNNNNNNNNNNNN | 1 |
| L | 2245 | hCoV-19/Belgium/ULG-10481/2020\|EPI_ISL_581590\|2020 | ···················· | ···················· | ···················· | ···················· | NNNNNNNNNNNNNNNNNNNN | NNNNNNNNNNNNNNNNNNNN | 1 |
| L | 2246 | hCoV-19/Belgium/ULG-10482/2020\|EPI_ISL_581591\|2020 | ···················· | ···················· | ···················· | ···················· | NNNNNNNNNNNNNNNNNNNN | NNNNNNNNNNNNNNNNNNNN | 1 |
| L | 2247 | hCoV-19/Belgium/ULG-10485/2020\|EPI_ISL_581592\|2020 | ···················· | ···················· | ···················· | ···················· | NNNNNNNNNNNNNNNNNNNN | NNNNNNNNNNNNNNNNNNNN | 1 |
| L | 2248 | hCoV-19/Belgium/ULG-10487/2020\|EPI_ISL_581593\|2020 | ···················· | ···················· | ···················· | ···················· | NNNNNNNNNNNNNNNNNNNN | NNNNNNNNNNNNNNNNNNNN | 1 |
| L | 2309 | hCoV-19/England/ALDP-9CC553/2020\|EPI_ISL_573113\|20 | ···················· | ···················· | ···················· | ···················· | NNNNNNNNNNNNNNNNNNNN | NNNNNNNNNNNNNNNNNNNN | 1 |
| L | 2310 | hCoV-19/England/ALDP-9CC5AE/2020\|EPI_ISL_573114\|20 | ···················· | ·A·················· | ···················· | ···················· | NNNNNNNNNNNNNNNNNNNN | NNNNNNNNNNNNNNNNNNNN | 1 |
| L | 2311 | hCoV-19/England/ALDP-9CC5BD/2020\|EPI_ISL_573115\|20 | ···················· | ···················· | ···················· | ···················· | NNNNNNNNNNNNNNNNNNNN | NNNNNNNNNNNNNNNNNNNN | 1 |
| L | 2317 | hCoV-19/England/ALDP-9CC702/2020\|EPI_ISL_573121\|20 | ···················· | ···················· | ···················· | ···················· | NNNNNNNNNNNNNNNNNNNN | NNNNNNNNNNNNNNNNNNNN | 1 |
| L | 2320 | hCoV-19/England/ALDP-9CCA09/2020\|EPI_ISL_573124\|20 | ···················· | ···················· | ···················· | ···················· | NNNNNNNNNNNNNNNNNNNN | NNNNNNNNNNNNNNNNNNNN | 1 |
| L | 2321 | hCoV-19/England/ALDP-9CCA63/2020\|EPI_ISL_573125\|20 | ···················· | ···················· | ···················· | ···················· | NNNNNNNNNNNNNNNNNNNN | NNNNNNNNNNNNNNNNNNNN | 1 |
| L | 2324 | hCoV-19/England/ALDP-9CCB8E/2020\|EPI_ISL_573128\|20 | ···················· | ·A·················· | ···················· | ···················· | NNNNNNNNNNNNNNNNNNNN | NNNNNNNNNNNNNNNNNNNN | 1 |
| L | 2325 | hCoV-19/England/ALDP-9CCBD9/2020\|EPI_ISL_573129\|20 | ···················· | ···················· | ···················· | ···················· | ···················· | NNNNN··············· | 1 |
| L | 2326 | hCoV-19/England/ALDP-9CCC4F/2020\|EPI_ISL_573130\|20 | ···················· | ·A·················· | ···················· | ···················· | NNNNNNNNNNNNNNNNNNNN | NNNNNNNNNNNNNNNNNNNN | 1 |
| L | 2327 | hCoV-19/England/ALDP-9CCC8B/2020\|EPI_ISL_573131\|20 | ···················· | ···················· | ···················· | ···················· | NNNNNNNNNNNNNNNNNNNN | NNNNNNNNNNNNNNNNNNNN | 1 |
| L | 2337 | hCoV-19/England/CAMB-1B5F57/2020\|EPI_ISL_573141\|20 | ···················· | ···················· | ···················· | ···················· | NNNNNNNNNNNNNNNNNNNN | NNNNNNNNNNNNNNNNNNNN | 1 |
| L | 2338 | hCoV-19/England/CAMB-1B5F66/2020\|EPI_ISL_573142\|20 | ···················· | ···················· | ···················· | ···················· | NNNNNNNNNNNNNNNNNNNN | NNNNNNNNNNNNNNNNNNNN | 1 |
| L | 2343 | hCoV-19/England/CAMB-1B6141/2020\|EPI_ISL_573147\|20 | ···················· | ···················· | ···················· | ···················· | NNNNNNNNNNNNNNNNNNNN | NNNNNNNNNNNNNNNNNNNN | 1 |
| L | 2344 | hCoV-19/England/CAMB-1B6150/2020\|EPI_ISL_573148\|20 | ···················· | ···················· | ···················· | ···················· | NNNNNNNNNNNNNNNNNNNN | NNNNNNNNNNNNNNNNNNNN | 1 |
| L | 2345 | hCoV-19/England/CAMB-1B616F/2020\|EPI_ISL_573149\|20 | ···················· | ···················· | ···················· | ···················· | NNNNNNNNNNNNNNNNNNNN | NNNNNNNNNNNNNNNNNNNN | 1 |
| L | 2349 | hCoV-19/England/CAMB-1B6202/2020\|EPI_ISL_573153\|20 | ···················· | ···················· | ···················· | ···················· | NNNNNNNNNNNNNNNNNNNN | NNNNNNNNNNNNNNNNNNNN | 1 |
| L | 2482 | hCoV-19/New Zealand/20VR2005/2020\|EPI_ISL_579200\|2 | ···················· | ···················· | ···················· | ···················· | NNNNNNNNNNNNNNNNNNNN | NNNNNNNNNNNNNNNNNNNN | 1 |
| L | 2485 | hCoV-19/New Zealand/20VR2010/2020\|EPI_ISL_579203\|2 | ···················· | ···················· | ···················· | ···················· | NNNNNNNNNNNNNNNNNNNN | NNNNNNNNNNNNNNNNNNNN | 1 |
| L | 2486 | hCoV-19/New Zealand/20VR2012/2020\|EPI_ISL_579204\|2 | ···················· | ···················· | ···················· | ···················· | NNNNNNNNNNNNNNNNNNNN | NNNNNNNNNNNNNNNNNNNN | 1 |
| L | 2532 | hCoV-19/Spain/MD-IBV-99007342/2020\|EPI_ISL_582826\| | ···················· | ···················· | ···················· | ···················· | NNNNNNNNNNNNNNNNNNNN | NNNNNNNNNNNNNNNNNNNN | 1 |
| L | 2533 | hCoV-19/Spain/MD-IBV-99007343/2020\|EPI_ISL_582827\| | ···················· | ···················· | ···················· | ···················· | NNNNNNN············· | NNNNNNNN······N····· | 1 |
| L | 2565 | hCoV-19/England/ALDP-9CC3C2/2020\|EPI_ISL_573104\|20 | ···················· | ·A·················· | ···················· | ···················· | NNNNNNNNNNNNNNNNNNNN | NNNNNNNNNNNNNNNNNNNN | 1 |
| L | 2566 | hCoV-19/England/ALDP-9CC3FF/2020\|EPI_ISL_573105\|20 | ···················· | ···················· | ···················· | ···················· | NNNNNNNNNNNNNNNNNNNN | NNNNNNNNNNNNNNNNNNNN | 1 |
| L | 2567 | hCoV-19/England/ALDP-9CC438/2020\|EPI_ISL_573106\|20 | ···················· | ···················· | ···················· | ···················· | ···················· | NNNNNNNNN··NNNNNNNNN | 1 |
| L | 2568 | hCoV-19/England/ALDP-9CC447/2020\|EPI_ISL_573107\|20 | ···················· | ···················· | ···················· | ···················· | NNNNNNNNNNNNNNNNNNNN | NNNNNNNNNNNNNNNNNNNN | 1 |
| L | 2569 | hCoV-19/England/ALDP-9CC456/2020\|EPI_ISL_573108\|20 | ···················· | ···················· | ···················· | ···················· | ···················· | NNNNN··············· | 1 |
| L | 2570 | hCoV-19/England/ALDP-9CC474/2020\|EPI_ISL_573109\|20 | ···················· | ···················· | ···················· | ···················· | NNNNNNNNNNNNNNNNNNNN | NNNNNNNNNNNNNNNNNNNN | 1 |
| L | 2573 | hCoV-19/England/ALDP-9CC508/2020\|EPI_ISL_573112\|20 | ···················· | ···················· | ···················· | ···················· | NNNNNNNNNNNNNNNNNNNN | NNNNNNNNNNNNNNNNNNNN | 1 |
| L | 2677 | hCoV-19/New Zealand/20VR1292/2020\|EPI_ISL_579123\|2 | ···················· | ···················· | ···················· | ···················· | NNNNNNNNNNNNNNNNNNNN | NNNNNNNNNNNNNNNNNNNN | 1 |
| L | 2678 | hCoV-19/New Zealand/20VR1760/2020\|EPI_ISL_579125\|2 | ···················· | ···················· | ···················· | ···················· | NNNNNNNNNNNNNNNNNNNN | NNNNNNNNNNNNNNNNNNNN | 1 |
| L | 2680 | hCoV-19/New Zealand/20VR1770/2020\|EPI_ISL_579127\|2 | ···················· | ···················· | ···················· | ···················· | NNNNNNNNNNNNNNNNNNNN | NNNNNNNNNNNNNNNNNNNN | 1 |
| L | 2701 | hCoV-19/New Zealand/20VR1850/2020\|EPI_ISL_579149\|2 | ···················· | ···················· | ···················· | ···················· | NNNNNNNNNNNNNNNNNNNN | NNNNNNNNNNNNNNNNNNNN | 1 |
| L | 3011 | hCoV-19/England/MILK-9917CA/2020\|EPI_ISL_576993\|20 | ···················· | ···················· | ···················· | ···················· | NNNNNNNNNNNNNNNNNNNN | NNNNNNNNNNNNNNNNNNNN | 1 |
| L | 3021 | hCoV-19/New Zealand/20VR1930/2020\|EPI_ISL_579166\|2 | ···················· | ···················· | ···················· | ···················· | NNNNNNNNNNNNNNNNNNNN | NNNNNNNNNNNNNNNNNNNN | 1 |
| L | 3023 | hCoV-19/New Zealand/20VR1935/2020\|EPI_ISL_579168\|2 | ···················· | ···················· | ···················· | ···················· | NNNNNNNNNNNNNNNNNNNN | NNNNNNNNNNNNNNNNNNNN | 1 |
| L | 3029 | hCoV-19/New Zealand/20VR1954/2020\|EPI_ISL_579175\|2 | ···················· | ···················· | ···················· | ···················· | NNNNNNNNNNNNNNNNNNNN | NNNNNNNNNNNNNNNNNNNN | 1 |
| L | 3190 | hCoV-19/England/CAMB-1B6475/2020\|EPI_ISL_576912\|20 | ···················· | ···················· | ···················· | ···················· | NNNNNNNNNNNNNNNNNNNN | NNNNNNNNNNNNNNNNNNNN | 1 |
| L | 3244 | hCoV-19/New Zealand/20VR1963/2020\|EPI_ISL_579182\|2 | ···················· | ···················· | ···················· | ···················· | NNNNNNNNNNNNNNNNNNNN | NNNNNNNNNNNNNNNNNNNN | 1 |
| L | 3247 | hCoV-19/New Zealand/20VR1978/2020\|EPI_ISL_579185\|2 | ···················· | ···················· | ···················· | ···················· | NNNNNNNNNNNNNNNNNNNN | NNNNNNNNNNNNNNNNNNNN | 1 |
| L | 3252 | hCoV-19/New Zealand/20VR1987/2020\|EPI_ISL_579190\|2 | ···················· | ···················· | ···················· | ···················· | NNNNNNNNNNNNNNNNNNNN | NNNNNNNNNNNNNNNNNNNN | 1 |
| L | 3414 | hCoV-19/Wales/PHWC-16CECB/2020\|EPI_ISL_574181\|2020 | ···················· | ···················· | ···················· | ···················· | NNNNNNNNNNNNNNNNNNNN | NNNNNNNNNNNNNNNNNNNN | 1 |
| L | 3415 | hCoV-19/Wales/PHWC-16CEE9/2020\|EPI_ISL_574182\|2020 | ···················· | ···················· | ···················· | ···················· | NNNNNNNNNNNN········ | ···················· | 1 |
| L | 3416 | hCoV-19/Wales/PHWC-16CEF8/2020\|EPI_ISL_574183\|2020 | ···················· | ···················· | ···················· | ···················· | NNNNNNNNNNNNNNNNNNNN | ···················· | 1 |
| L | 3423 | hCoV-19/Wales/PHWC-16CF9B/2020\|EPI_ISL_574190\|2020 | ···················· | ···················· | ···················· | ···················· | NNNNNNNNNNNNNNNNNNNN | NNNNNNNNNNNNNNNNNNNN | 1 |
| L | 3428 | hCoV-19/Wales/PHWC-16CFF5/2020\|EPI_ISL_574195\|2020 | ···················· | ···················· | ···················· | ···················· | NNNNNNNNNNNNNNNNNNNN | NNNNNNNNNNNNNNNNNNNN | 1 |
| L | 3433 | hCoV-19/Wales/PHWC-16D2CE/2020\|EPI_ISL_574222\|2020 | ···················· | ···················· | ···················· | ···················· | NNNNN··············· | ···················· | 1 |
| L | 3589 | hCoV-19/Wales/PHWC-16D158/2020\|EPI_ISL_574206\|2020 | ···················· | ···················· | ···················· | ···················· | NNNNNNNNNNNNNNNNNNNN | NNNNNNNNNNNNNNN··NNN | 1 |
| L | 3592 | hCoV-19/Wales/PHWC-16D1C1/2020\|EPI_ISL_574209\|2020 | ···················· | ···················· | ···················· | ···················· | NNNNNNNNNNNNNNNNNNNN | NNNNNNNNNNNNNNNNNNNN | 1 |
| L | 3594 | hCoV-19/Wales/PHWC-16D1FE/2020\|EPI_ISL_574211\|2020 | ···················· | ···················· | ···················· | ···················· | NNNNNNNNNNNNNNNNNNNN | NNNNNNNNNNNNNNN····· | 1 |
| L | 3595 | hCoV-19/Wales/PHWC-16D20A/2020\|EPI_ISL_574212\|2020 | ···················· | ···················· | ···················· | ···················· | NNNN················ | ···················· | 1 |
| L | 3596 | hCoV-19/Wales/PHWC-16D219/2020\|EPI_ISL_574213\|2020 | ···················· | ···················· | ···················· | ···················· | NNNN················ | ···················· | 1 |
| L | 3597 | hCoV-19/Wales/PHWC-16D228/2020\|EPI_ISL_574214\|2020 | ···················· | ···················· | ···················· | ···················· | ···················· | NNNNNNNNNNNNNNNNNNNN | 1 |
| L | 3601 | hCoV-19/Wales/PHWC-16D264/2020\|EPI_ISL_574218\|2020 | ···················· | ···················· | ···················· | ···················· | NNNNNNNNNNNNNN······ | ···················· | 1 |
| L | 3608 | hCoV-19/Wales/PHWC-16D307/2020\|EPI_ISL_574226\|2020 | ···················· | ···················· | ···················· | ···················· | NNNNNNNNNNNNNNNNNNNN | NNNNNNNNNNNNNNNNNNNN | 1 |
| L | 3610 | hCoV-19/Wales/PHWC-16D370/2020\|EPI_ISL_574228\|2020 | ···················· | ···················· | ···················· | ···················· | NNNNNNNNNNNNNNNNNNNN | ···················· | 1 |
| L | 3612 | hCoV-19/Wales/PHWC-16D39E/2020\|EPI_ISL_574230\|2020 | ···················· | ···················· | ···················· | ···················· | NNNNNNNNNNNNNNNNNNNN | ···················· | 1 |
| L | 3617 | hCoV-19/Wales/PHWC-16D413/2020\|EPI_ISL_574235\|2020 | ···················· | ···················· | ···················· | ···················· | NNNNNNNNNNNNNNNNNNNN | NNNNNNNNNNNNNNNNNNNN | 1 |
| L | 3623 | hCoV-19/Wales/PHWC-16D49B/2020\|EPI_ISL_574241\|2020 | ···················· | ···················· | ···················· | ···················· | NNNNNNNNNNNNN······· | ···················· | 1 |
| L | 3626 | hCoV-19/Wales/PHWC-16D4C8/2020\|EPI_ISL_574244\|2020 | ···················· | ···················· | ···················· | ···················· | NNNNNNNNNNNNNN······ | ···················· | 1 |
| L | 3630 | hCoV-19/Wales/PHWC-16D510/2020\|EPI_ISL_574248\|2020 | ···················· | ···················· | ···················· | ···················· | NNNNNNNNNNNNNNNNNNNN | NNNNNNNNNNNNNNNNNNNN | 1 |

| | | | | | | | | | |
|---|---|---|---|---|---|---|---|---|---|
| L | 3631 | hCoV-19/Wales/PHWC-16D54D/2020\|EPI_ISL_574249\|2020 | ···················· | ···················· | ···················· | ···················· | NNNNNN·············· | ···················· | 1 |
| L | 3637 | hCoV-19/Wales/PHWC-16D5A7/2020\|EPI_ISL_574255\|2020 | ···················· | ···················· | ···················· | ···················· | NNNNNNNNNNNNNNNNNNNN | ···················· | 1 |
| L | 3750 | hCoV-19/England/BIRM-63C9E/2020\|EPI_ISL_576877\|202 | ···················· | ···················· | ···················· | ···················· | NNNNNNNNNNNNNNNNNNNN | NNNNNNNNNNNNNNNNNNNN | 1 |
| L | 3751 | hCoV-19/England/BIRM-63D7D/2020\|EPI_ISL_576882\|202 | ···················· | ···················· | ···················· | ···················· | NNNNNNNNNNNNNNNNNNNN | NNNNNNNNNNNNNNNNNNNN | 1 |
| L | 3761 | hCoV-19/England/CAMB-1B62A8/2020\|EPI_ISL_576897\|20 | ···················· | ···················· | ···················· | ···················· | NNNNNNNNNNNNNNNNNNNN | NNNNNNNNNNNNNNNNNNNN | 1 |
| L | 3786 | hCoV-19/New Zealand/20CV0325/2020\|EPI_ISL_579090\|2 | ···················· | ···················· | ···················· | ···················· | NNNNNNNNNNNNNNNNNNNN | NNNNNNNNNNNNNNNNNNNN | 1 |
| L | 3787 | hCoV-19/New Zealand/20CV0332/2020\|EPI_ISL_579092\|2 | ···················· | ·A·················· | ···················· | ···················· | NNNNNNNNNNNNNNNNNNNN | NNNNNNNNNNNNNNNNNNNN | 1 |
| L | 3901 | hCoV-19/Wales/PHWC-16C58A/2020\|EPI_ISL_574067\|2020 | ···················· | ···················· | ···················· | ···················· | NNNNNNNNNNNNNNNNNNNN | NNNNNNNNNNNNNNNNNNNN | 1 |
| L | 3903 | hCoV-19/Wales/PHWC-16C5A8/2020\|EPI_ISL_574069\|2020 | ···················· | ···················· | ···················· | ···················· | NNNNNNNNNNNNNNNNNNNN | ···················· | 1 |
| L | 3904 | hCoV-19/Wales/PHWC-16D0D3/2020\|EPI_ISL_574200\|2020 | ···················· | ···················· | ···················· | ···················· | NNNNNNNNNNNNNNNNNNNN | NNNNNNNNNNNNNNNNNNNN | 1 |
| L | 4149 | hCoV-19/Wales/PHWC-16C3CC/2020\|EPI_ISL_574050\|2020 | ···················· | ···················· | ···················· | ···················· | NNNNNNNNNNNNNNNNNNNN | ···················· | 1 |
| L | 4150 | hCoV-19/Wales/PHWC-16C423/2020\|EPI_ISL_574055\|2020 | ···················· | ···················· | ···················· | ···················· | NNNNNNNNNNNNNNNNNNNN | ···················· | 1 |
| L | 4153 | hCoV-19/Wales/PHWC-16C5B7/2020\|EPI_ISL_574070\|2020 | ···················· | ···················· | ···················· | ···················· | NNN·NNNNNNNNNNNNNNNN | ···················· | 1 |
| L | 4162 | hCoV-19/Wales/PHWC-16C65A/2020\|EPI_ISL_574079\|2020 | ···················· | ···················· | ···················· | ···················· | NNNNNNNNNNNNNNNNNNNN | NNNNNNNNNNNNNNNNNNNN | 1 |
| L | 4180 | hCoV-19/Wales/PHWC-16C7FD/2020\|EPI_ISL_574097\|2020 | ···················· | ···················· | ···················· | ···················· | NNN················· | NNNNNNNNNNNN········ | 1 |
| L | 4189 | hCoV-19/Wales/PHWC-16C8BE/2020\|EPI_ISL_574106\|2020 | ···················· | ···················· | ···················· | ···················· | NNNNNNNNNNNN········ | ···················· | 1 |
| L | 4195 | hCoV-19/Wales/PHWC-16C933/2020\|EPI_ISL_574112\|2020 | ···················· | ···················· | ···················· | ···················· | NNNNNNNNNNNNNNNNN··· | ···················· | 1 |
| L | 4201 | hCoV-19/Wales/PHWC-16C9D9/2020\|EPI_ISL_574118\|2020 | ···················· | ···················· | ···················· | ···················· | NNNNNNNNNNNNNNNNNNNN | ········NNNNNNNNNNNN | 1 |
| L | 4212 | hCoV-19/Wales/PHWC-16CA9A/2020\|EPI_ISL_574129\|2020 | ···················· | ···················· | ···················· | ···················· | NNNNNNNNNNNNNNNNNNNN | NNNNNNNNNNNNNNNNNNNN | 1 |
| L | 4213 | hCoV-19/Wales/PHWC-16CAA9/2020\|EPI_ISL_574130\|2020 | ···················· | ···················· | ···················· | ···················· | NNNNNNNNNNNNNNNNNNNN | NNNN················ | 1 |
| L | 4214 | hCoV-19/Wales/PHWC-16CAB8/2020\|EPI_ISL_574131\|2020 | ···················· | ···················· | ···················· | ···················· | NNNNNNNNNNNNNNNNNNNN | ···················· | 1 |
| L | 4362 | hCoV-19/Wales/PHWC-16CBF1/2020\|EPI_ISL_574147\|2020 | ···················· | ···················· | ···················· | ···················· | NNNN················ | ···················· | 1 |
| L | 4369 | hCoV-19/Wales/PHWC-16CC94/2020\|EPI_ISL_574154\|2020 | ···················· | ···················· | ···················· | ···················· | NNNN····NNNNNNNNNNNN | ···················· | 1 |
| L | 4370 | hCoV-19/Wales/PHWC-16CCA3/2020\|EPI_ISL_574155\|2020 | ···················· | ···················· | ···················· | ···················· | NNNNNNNNNNNNNNNNNNNN | NNNNNNNNNNNNNNNNNNNN | 1 |
| L | 4371 | hCoV-19/Wales/PHWC-16CCB2/2020\|EPI_ISL_574156\|2020 | ···················· | ···················· | ···················· | ···················· | NNNN················ | ···················· | 1 |
| L | 4375 | hCoV-19/Wales/PHWC-16CD28/2020\|EPI_ISL_574160\|2020 | ···················· | ···················· | ···················· | ···················· | NNNNNNNNNNNNNNNNNNNN | ···················· | 1 |
| L | 4377 | hCoV-19/Wales/PHWC-16CDA0/2020\|EPI_ISL_574166\|2020 | ···················· | ···················· | ···················· | ···················· | NNNNNNNNNNNNNNNNNNNN | NNNNNNNNNNNNNNNNNNNN | 1 |
| L | 4516 | hCoV-19/USA/WI-WSLH-200143/2020\|EPI_ISL_578370\|202 | ···················· | ···················· | ···················· | ···················· | NNNNNNNNNNNNNNNNNNNN | NNNNNNNNNNNNNNNNNNNN | 1 |
| L | 4517 | hCoV-19/USA/WI-WSLH-200144/2020\|EPI_ISL_578371\|202 | ···················· | ···················· | ···················· | ···················· | NNNNNNNNNNNNNNNNNNNN | NNNNNNNNNNNNNNNNNNNN | 1 |
| L | 4518 | hCoV-19/USA/WI-WSLH-200145/2020\|EPI_ISL_578372\|202 | ···················· | ···················· | ···················· | ···················· | NNNNNNNNNNNNNNNNNNNN | NNNNNNNNNNNNNNNNNNNN | 1 |
| L | 4519 | hCoV-19/USA/WI-WSLH-200146/2020\|EPI_ISL_578373\|202 | ···················· | ···················· | ···················· | ···················· | NNNNNNNNNNNNNNNNNNNN | NNNNNNNNNNNNNNNNNNNN | 1 |
| L | 4520 | hCoV-19/USA/WI-WSLH-200147/2020\|EPI_ISL_578374\|202 | ···················· | ···················· | ···················· | ···················· | NNNNNNNNNNNNNNNNNNNN | NNNNNNNNNNNNNNNNNNNN | 1 |
| L | 4521 | hCoV-19/USA/WI-WSLH-200148/2020\|EPI_ISL_578375\|202 | ···················· | ···················· | ···················· | ···················· | NNNNNNNNNNNNNNNNNNNN | NNNNNNNNNNNNNNNNNNNN | 1 |
| L | 4522 | hCoV-19/USA/WI-WSLH-200149/2020\|EPI_ISL_578376\|202 | ···················· | ···················· | ···················· | ···················· | NNNNNNNNNNNNNNNNNNNN | NNNNNNNNNNNNNNNNNNNN | 1 |
| L | 4523 | hCoV-19/USA/WI-WSLH-200150/2020\|EPI_ISL_578377\|202 | ···················· | ···················· | ···················· | ···················· | NNNNNNNNNNNNNNNNNNNN | NNNNNNNNNNNNNNNNNNNN | 1 |
| L | 4524 | hCoV-19/USA/WI-WSLH-200151/2020\|EPI_ISL_578378\|202 | ···················· | ···················· | ···················· | ···················· | NNNNNNNNNNNNNNNNNNNN | NNNNNNNNNNNNNNNNNNNN | 1 |
| L | 4525 | hCoV-19/USA/WI-WSLH-200152/2020\|EPI_ISL_578379\|202 | ···················· | ···················· | ···················· | ···················· | NNNNNNNNNNNNNNNNNNNN | NNNNNNNNNNNNNNNNNNNN | 1 |
| L | 4528 | hCoV-19/USA/WI-WSLH-200155/2020\|EPI_ISL_578382\|202 | ···················· | ···················· | ···················· | ···················· | NNNNNNNNNNNNNNNNNNNN | NNNNNNNNNNNNNNNNNNNN | 1 |
| L | 4529 | hCoV-19/USA/WI-WSLH-200156/2020\|EPI_ISL_578383\|202 | ···················· | ···················· | ···················· | ···················· | NNNNNNNNNNNNNNNNNNNN | NNNNNNNNNNNNNNNNNNNN | 1 |
| L | 4530 | hCoV-19/USA/WI-WSLH-200157/2020\|EPI_ISL_578384\|202 | ···················· | ···················· | ···················· | ···················· | NNNNNNNNNNNNNNNNNNNN | NNNNNNNNNNNNNNNNNNNN | 1 |
| L | 4531 | hCoV-19/USA/WI-WSLH-200158/2020\|EPI_ISL_578385\|202 | ···················· | ···················· | ···················· | ···················· | NNNNNNNNNNNNNNNNNNNN | NNNNNNNNNNNNNNNNNNNN | 1 |
| L | 4535 | hCoV-19/USA/WI-WSLH-200163/2020\|EPI_ISL_578389\|202 | ···················· | ···················· | ···················· | ···················· | NNNNNNNNNNNNNNNNNNNN | NNNNNNNNNNNNNNNNNNNN | 1 |
| L | 4536 | hCoV-19/USA/WI-WSLH-200164/2020\|EPI_ISL_578390\|202 | ···················· | ···················· | ···················· | ···················· | NNNNNNNNNNNNNNNNNNNN | NNNNNNNNNNNNNNNNNNNN | 1 |
| L | 4537 | hCoV-19/USA/WI-WSLH-200165/2020\|EPI_ISL_578391\|202 | ···················· | ···················· | ···················· | ···················· | NNNNNNNNNNNNNNNNNNNN | NNNNNNNNNNNNNNNNNNNN | 1 |
| L | 4538 | hCoV-19/USA/WI-WSLH-200166/2020\|EPI_ISL_578392\|202 | ···················· | ···················· | ···················· | ···················· | NNNNNNNNNNNNNNNNNNNN | NNNNNNNNNNNNNNNNNNNN | 1 |
| L | 4539 | hCoV-19/USA/WI-WSLH-200167/2020\|EPI_ISL_578393\|202 | ···················· | ···················· | ···················· | ···················· | NNNNNNNNNNNNNNNNNNNN | NNNNNNNNNNNNNNNNNNNN | 1 |
| L | 4540 | hCoV-19/USA/WI-WSLH-200168/2020\|EPI_ISL_578394\|202 | ···················· | ···················· | ···················· | ···················· | NNNNNNNNNNNNNNNNNNNN | NNNNNNNNNNNNNNNNNNNN | 1 |
| L | 4541 | hCoV-19/USA/WI-WSLH-200169/2020\|EPI_ISL_578395\|202 | ···················· | ···················· | ···················· | ···················· | NNNNNNNNNNNNNNNNNNNN | NNNNNNNNNNNNNNNNNNNN | 1 |
| L | 4542 | hCoV-19/USA/WI-WSLH-200170/2020\|EPI_ISL_578396\|202 | ···················· | ···················· | ···················· | ···················· | NNNNNNNNNNNNNNNNNNNN | NNNNNNNNNNNNNNNNNNNN | 1 |
| L | 4545 | hCoV-19/USA/WI-WSLH-200173/2020\|EPI_ISL_578399\|202 | ···················· | ···················· | ···················· | ···················· | NNNNNNNNNNNNNNNNNNNN | NNNNNNNNNNNNNNNNNNNN | 1 |
| L | 4607 | hCoV-19/Switzerland/BS-42393291/2020\|EPI_ISL_58195 | ···················· | ···················· | ···················· | ···················· | NNNNNNNNNNNNNNNNNNNN | NNNNNNNNNNNNNNNNNNNN | 1 |
| L | 4608 | hCoV-19/Switzerland/BS-42394001/2020\|EPI_ISL_58195 | ···················· | ···················· | ···················· | ···················· | NNNNNNNNNNNNNNNNNNNN | NNNNNNNNNNNNNNNNNNNN | 1 |
| L | 4633 | hCoV-19/USA/VA-DCLS-1275/2020\|EPI_ISL_572291\|2020- | ···················· | ···················· | ···················· | ···················· | ··NNNNNNNNNNNNNNNNNN | ···················· | 1 |
| L | 4652 | hCoV-19/England/MILK-9919C4/2020\|EPI_ISL_577010\|20 | ···················· | ···················· | ···················· | ···················· | NNNNNNNNNNNNNNNNNNNN | NNNNNNNNNNNNNNNNNNNN | 1 |
| L | 4654 | hCoV-19/England/MILK-9919E2/2020\|EPI_ISL_577012\|20 | ···················· | ···················· | ···················· | ···················· | NNNNNNNNNNNNNNNNNNNN | NNNNNNNNNNNNNNNNNNNN | 1 |
| L | 4656 | hCoV-19/England/MILK-991A2B/2020\|EPI_ISL_577014\|20 | ···················· | ···················· | ···················· | ···················· | NNNNNNNNNNNNNNNNNNNN | NNNNNNNNNNNNNNNNNNNN | 1 |
| L | 4671 | hCoV-19/England/MILK-991BFB/2020\|EPI_ISL_577031\|20 | ···················· | ···················· | ···················· | ···················· | NNNNNNNNNNNNNNNNNNNN | NNNNNNNNNNNNNNNNNNNN | 1 |
| L | 4676 | hCoV-19/England/MILK-991C70/2020\|EPI_ISL_577036\|20 | ···················· | ···················· | ···················· | ···················· | NNNNNNNNNNNNNNNNNNNN | NNNNNNNNNNNNNNNNNNNN | 1 |
| L | 4684 | hCoV-19/Ireland/KK-NVRL-73IRL21709/2020\|EPI_ISL_57 | ···················· | ·A·················· | ···················· | ···················· | NNNNNNNNNNNNNNNNNNNN | NNNNNNNNNNNNNNNNNNNN | 1 |
| L | 4700 | hCoV-19/Ireland/WD-NVRL-72IRL52688/2020\|EPI_ISL_57 | ···················· | ·A·················· | ···················· | ···················· | NNNNNNNNNNNNNNNNNNNN | NNNNNNNNNNNNNNNNNNNN | 1 |
| L | 4701 | hCoV-19/Ireland/WD-NVRL-72IRL52708/2020\|EPI_ISL_57 | ···················· | ·A·················· | ···················· | ···················· | NNNNNNNNNNNNNNNNNNNN | NNNNNNNNNNNNNNNNNNNN | 1 |
| L | 4705 | hCoV-19/Ireland/WW-NVRL-73IRL23922/2020\|EPI_ISL_57 | ···················· | ···················· | ···················· | ···················· | NNNNNNNNNNNNNNNNNNNN | NNNNNNNNNNNNNNNNNNNN | 1 |
| L | 4707 | hCoV-19/Ireland/WW-NVRL-73IRL29878/2020\|EPI_ISL_57 | ···················· | ···················· | ···················· | ···················· | NNNNNNNNNNNNNNNNNNNN | NNNNNNNNNNNNNNNNNNNN | 1 |
| L | 4713 | hCoV-19/USA/WI-WSLH-200105/2020\|EPI_ISL_578337\|202 | ···················· | ···················· | ···················· | ···················· | NNNNNNNNNNNNNNNNNNNN | NNNNNNNNNNNNNNNNNNNN | 1 |
| L | 4716 | hCoV-19/USA/WI-WSLH-200109/2020\|EPI_ISL_578340\|202 | ···················· | ···················· | ···················· | ···················· | NNNNNNNNNNNNNNNNNNNN | NNNNNNNNNNNNNNNNNNNN | 1 |
| L | 4719 | hCoV-19/USA/WI-WSLH-200112/2020\|EPI_ISL_578343\|202 | ···················· | ···················· | ···················· | ···················· | NNNNNNNNNNNNNNNNNNNN | NNNNNNNNNNNNNNNNNNNN | 1 |
| L | 4721 | hCoV-19/USA/WI-WSLH-200115/2020\|EPI_ISL_578345\|202 | ···················· | ···················· | ···················· | ···················· | NNNNNNNNNNNNNNNNNNNN | NNNNNNNNNNNNNNNNNNNN | 1 |
| L | 4722 | hCoV-19/USA/WI-WSLH-200116/2020\|EPI_ISL_578346\|202 | ···················· | ···················· | ···················· | ···················· | NNNNNNNNNNNNNNNNNNNN | NNNNNNNNNNNNNNNNNNNN | 1 |
| L | 4723 | hCoV-19/USA/WI-WSLH-200117/2020\|EPI_ISL_578347\|202 | ···················· | ···················· | ···················· | ···················· | NNNNNNNNNNNNNNNNNNNN | NNNNNNNNNNNNNNNNNNNN | 1 |
| L | 4726 | hCoV-19/USA/WI-WSLH-200121/2020\|EPI_ISL_578350\|202 | ···················· | ···················· | ···················· | ···················· | NNNNNNNNNNNNNNNNNNNN | NNNNNNNNNNNNNNNNNNNN | 1 |
| L | 4727 | hCoV-19/USA/WI-WSLH-200122/2020\|EPI_ISL_578351\|202 | ···················· | ···················· | ···················· | ···················· | NNNNNNNNNNNNNNNNNNNN | NNNNNNNNNNNNNNNNNNNN | 1 |
| L | 4728 | hCoV-19/USA/WI-WSLH-200123/2020\|EPI_ISL_578352\|202 | ···················· | ···················· | ···················· | ···················· | NNNNNNNNNNNNNNNNNNNN | NNNNNNNNNNNNNNNNNNNN | 1 |
| L | 4729 | hCoV-19/USA/WI-WSLH-200124/2020\|EPI_ISL_578353\|202 | ···················· | ···················· | ···················· | ···················· | NNNNNNNNNNNNNNNNNNNN | NNNNNNNNNNNNNNNNNNNN | 1 |
| L | 4731 | hCoV-19/USA/WI-WSLH-200126/2020\|EPI_ISL_578355\|202 | ···················· | ···················· | ···················· | ···················· | NNNNNNNNNNNNNNNNNNNN | NNNNNNNNNNNNNNNNNNNN | 1 |
| L | 4732 | hCoV-19/USA/WI-WSLH-200127/2020\|EPI_ISL_578356\|202 | ···················· | ···················· | ···················· | ···················· | NNNNNNNNNNNNNNNNNNNN | NNNNNNNNNNNNNNNNNNNN | 1 |
| L | 4733 | hCoV-19/USA/WI-WSLH-200129/2020\|EPI_ISL_578357\|202 | ···················· | ···················· | ···················· | ···················· | NNNNNNNNNNNNNNNNNNNN | NNNNNNNNNNNNNNNNNNNN | 1 |
| L | 4734 | hCoV-19/USA/WI-WSLH-200130/2020\|EPI_ISL_578358\|202 | ···················· | ···················· | ···················· | ···················· | NNNNNNNNNNNNNNNNNNNN | NNNNNNNNNNNNNNNNNNNN | 1 |
| L | 4735 | hCoV-19/USA/WI-WSLH-200131/2020\|EPI_ISL_578359\|202 | ···················· | ···················· | ···················· | ···················· | NNNNNNNNNNNNNNNNNNNN | NNNNNNNNNNNNNNNNNNNN | 1 |
| L | 4736 | hCoV-19/USA/WI-WSLH-200132/2020\|EPI_ISL_578360\|202 | ···················· | ···················· | ···················· | ···················· | NNNNNNNNNNNNNNNNNNNN | NNNNNNNNNNNNNNNNNNNN | 1 |
| L | 4738 | hCoV-19/USA/WI-WSLH-200134/2020\|EPI_ISL_578362\|202 | ···················· | ···················· | ···················· | ···················· | NNNNNNNNNNNNNNNNNNNN | NNNNNNNNNNNNNNNNNNNN | 1 |

| | | | | | | | | | |
|---|---|---|---|---|---|---|---|---|---|
| L | 4739 | hCoV-19/USA/WI-WSLH-200135/2020\|EPI_ISL_578363\|202 | ···················· | ···················· | ···················· | ···················· | NNNNNNNNNNNNNNNNNNNN | NNNNNNNNNNNNNNNNNNNN | 1 |
| L | 4740 | hCoV-19/USA/WI-WSLH-200136/2020\|EPI_ISL_578364\|202 | ···················· | ···················· | ···················· | ···················· | NNNNNNNNNNNNNNNNNNNN | NNNNNNNNNNNNNNNNNNNN | 1 |
| L | 4741 | hCoV-19/USA/WI-WSLH-200137/2020\|EPI_ISL_578365\|202 | ···················· | ···················· | ···················· | ···················· | NNNNNNNNNNNNNNNNNNNN | NNNNNNNNNNNNNNNNNNNN | 1 |
| L | 4742 | hCoV-19/USA/WI-WSLH-200139/2020\|EPI_ISL_578366\|202 | ···················· | ···················· | ···················· | ···················· | NNNNNNNNNNNNNNNNNNNN | NNNNNNNNNNNNNNNNNNNN | 1 |
| L | 4743 | hCoV-19/USA/WI-WSLH-200140/2020\|EPI_ISL_578367\|202 | ···················· | ···················· | ···················· | ···················· | NNNNNNNNNNNNNNNNNNNN | NNNNNNNNNNNNNNNNNNNN | 1 |
| L | 4744 | hCoV-19/USA/WI-WSLH-200141/2020\|EPI_ISL_578368\|202 | ···················· | ···················· | ···················· | ···················· | NNNNNNNNNNNNNNNNNNNN | NNNNNNNNNNNNNNNNNNNN | 1 |
| L | 4745 | hCoV-19/USA/WI-WSLH-200142/2020\|EPI_ISL_578369\|202 | ···················· | ···················· | ···················· | ···················· | NNNNNNNNNNNNNNNNNNNN | NNNNNNNNNNNNNNNNNNNN | 1 |
| L | 4825 | hCoV-19/Switzerland/BS-42374717/2020\|EPI_ISL_58193 | ···················· | ···················· | ···················· | ···················· | NNNNNNNNNNNNNNNNNNNN | NNNNNNNNNNNNNNNNN··· | 1 |
| L | 4827 | hCoV-19/Switzerland/BS-42375489/2020\|EPI_ISL_58193 | ···················· | ···················· | ···················· | ···················· | NNNNNNNNNNNNNNNNNNNN | NNNNNNNNNNNNNNNNNNNN | 1 |
| L | 4828 | hCoV-19/Switzerland/BS-42376604/2020\|EPI_ISL_58193 | ···················· | ···················· | ···················· | ···················· | NNNNNNNNNNNNNNNNNNNN | ···················· | 1 |
| L | 4829 | hCoV-19/Switzerland/BS-42378086/2020\|EPI_ISL_58193 | ···················· | ···················· | ···················· | ···················· | NNNNNNNNNNNNNNNNNNNN | ···················· | 1 |
| L | 4830 | hCoV-19/Switzerland/BS-42378851/2020\|EPI_ISL_58193 | ···················· | ···················· | ···················· | ···················· | NNNNNNNNNNNNNNNNNNNN | NNNNN··············· | 1 |
| L | 4831 | hCoV-19/Switzerland/BS-42379000/2020\|EPI_ISL_58193 | ···················· | ···················· | ···················· | ···················· | NNNNNNNNNNNNNNNNNNNN | NNNNNNNNNNNNNNNNNNNN | 1 |
| L | 4832 | hCoV-19/Switzerland/BS-42382638/2020\|EPI_ISL_58193 | ···················· | ···················· | ···················· | ···················· | NNNNNNNNNNNNNNNNNNNN | NNNNNNNNNNNNNNNNNNNN | 1 |
| L | 4833 | hCoV-19/Switzerland/BL-42384079/2020\|EPI_ISL_58193 | ···········T········ | ···················· | ···················· | ···················· | NNNNNNNNNNNNNNNNNNNN | NNNNNNNNNNNNNNNNNNNN | 1 |
| L | 4834 | hCoV-19/Switzerland/BL-42384084/2020\|EPI_ISL_58193 | ···········T········ | ···················· | ···················· | ···················· | NNNNNNNNNNNNNNNNNNNN | NNNNNNNNNNNNNNNNNNNN | 1 |
| L | 4835 | hCoV-19/Switzerland/BS-42384553/2020\|EPI_ISL_58194 | ···················· | ···················· | ···················· | ···················· | NNNNNNNNNNNNNNNNNNNN | NNNNNNNNNNNNNNNNNNNN | 1 |
| L | 4836 | hCoV-19/Switzerland/BS-42385186/2020\|EPI_ISL_58194 | ···················· | ···················· | ···················· | ···················· | NNNNNNNNNNNNNNNNNNNN | NNNNNNNNNNNNNNNNNNNN | 1 |
| L | 4837 | hCoV-19/Switzerland/BS-42386285/2020\|EPI_ISL_58194 | ···················· | ···················· | ···················· | ···················· | NNNNNNNNNNNNNNNNNNNN | NNNNNNNNNNNNNNNNNNNN | 1 |
| L | 4838 | hCoV-19/Switzerland/AG-42387010/2020\|EPI_ISL_58194 | ···················· | ···················· | ···················· | ···················· | NNNNNNNNNNNNNNNNNNNN | ···················· | 1 |
| L | 4840 | hCoV-19/Switzerland/BS-42388921/2020\|EPI_ISL_58194 | ···················· | ···················· | ···················· | ···················· | NNNNNNNNNNNNNNNNNNNN | NNNNNNNNNNNNNNNNNNNN | 1 |
| L | 4842 | hCoV-19/Switzerland/BS-42389973/2020\|EPI_ISL_58194 | ···················· | ···················· | ···················· | ···················· | NNNNNNNNNNNNNNNNNNNN | NNNNNNNNNNNNNNNNNNNN | 1 |
| L | 4843 | hCoV-19/Switzerland/BS-42390248/2020\|EPI_ISL_58194 | ···················· | ···················· | ···················· | ···················· | NNNNNNNNNNNNNNNNNNNN | NNNNNNNNNNNNNNNNNNNN | 1 |
| L | 4844 | hCoV-19/Switzerland/BS-42390260/2020\|EPI_ISL_58194 | ···················· | ···················· | ···················· | ···················· | NNNNNNNNNNNNNNNNNNNN | NNNNNNNNNNNNNNNNNNNN | 1 |
| L | 4846 | hCoV-19/Switzerland/BS-42392007/2020\|EPI_ISL_58195 | ···················· | ···················· | ···················· | ···················· | ·NNNNNNNNNNNNNNNNNNN | NNNNNNNNN··········· | 1 |
| L | 4848 | hCoV-19/Switzerland/BS-42395177/2020\|EPI_ISL_58195 | ···················· | ···················· | ···················· | ···················· | ·NNNNNNNN··········· | ···················· | 1 |
| L | 4850 | hCoV-19/Switzerland/BS-42395355/2020\|EPI_ISL_58195 | ···················· | ···················· | ···················· | ···················· | NNNNNNNNNNNNNNNNNNNN | ···················· | 1 |
| L | 4853 | hCoV-19/Switzerland/BS-65254283/2020\|EPI_ISL_58196 | ···················· | ···················· | ···················· | ···················· | NNNNNNNNNNNNNNNNNNNN | NNNNNNNNNNNNNNNNNNNN | 1 |
| L | 4854 | hCoV-19/Switzerland/BL-65254607/2020\|EPI_ISL_58196 | ···········T········ | ···················· | ···················· | ···················· | NNNNNNNNNNNNNNNNNNNN | ···················· | 1 |
| L | 4855 | hCoV-19/Switzerland/BS-65254979/2020\|EPI_ISL_58196 | ···················· | ···················· | ···················· | ···················· | NNNNNNN············· | ···················· | 1 |
| L | 4858 | hCoV-19/Switzerland/BL-42402436/2020\|EPI_ISL_58197 | ···················· | ···················· | ···················· | ···················· | NNNNNNNNNNNNNNNNNNNN | NNNNNNNNNNNNNNNNNNNN | 1 |
| L | 4860 | hCoV-19/Switzerland/BS-42404943/2020\|EPI_ISL_58198 | ···················· | ···················· | ···················· | ···················· | NNNNN··············· | NNNNN··············· | 1 |
| L | 4861 | hCoV-19/Switzerland/BS-42406439/2020\|EPI_ISL_58198 | ···················· | ·A·················· | ···················· | ···················· | NNNNNNNNNNNNNNNNNNNN | NNNNNNNNNNNNNNNNNNNN | 1 |
| L | 4862 | hCoV-19/Switzerland/BS-42408481/2020\|EPI_ISL_58198 | ···················· | ···G················ | ···················· | ···················· | NNNNNNNNNNNNNNNNNNNN | NNNNNNNNNNNNNNNNNNNN | 1 |
| L | 4863 | hCoV-19/Switzerland/BL-42412116/2020\|EPI_ISL_58198 | ···················· | ···················· | ···················· | ···················· | NNNNNNNNNNNNNNNNNNNN | NNNNNNNNNNNNNNNNNNNN | 1 |
| L | 4864 | hCoV-19/Switzerland/BS-42412257/2020\|EPI_ISL_58198 | ···················· | ···················· | ···················· | ···················· | NNNNNNNNNNNNNNNNNNNN | NNNNNNNNNNNNNNNNNNNN | 1 |
| L | 5064 | hCoV-19/Ireland/D-NVRL-20ILR05741/2020\|EPI_ISL_578 | ···················· | ·A·················· | ···················· | ···················· | NNNNNNNNNNNNNNNNNNNN | NNNNNNNNNNNNNNNNNNNN | 1 |
| L | 5078 | hCoV-19/USA/WI-WSLH-200174/2020\|EPI_ISL_578400\|202 | ···················· | ···················· | ···················· | ···················· | NNNNNNNNNNNNNNNNNNNN | NNNNNNNNNNNNNNNNNNNN | 1 |
| L | 5080 | hCoV-19/USA/WI-WSLH-200176/2020\|EPI_ISL_578402\|202 | ···················· | ···················· | ···················· | ···················· | NNNNNNNNNNNNNNNNNNNN | NNNNNNNNNNNNNNNNNNNN | 1 |
| L | 5081 | hCoV-19/USA/WI-WSLH-200177/2020\|EPI_ISL_578403\|202 | ···················· | ···················· | ···················· | ···················· | NNNNNNNNNNNNNNNNNNNN | NNNNNNNNNNNNNNNNNNNN | 1 |
| L | 5180 | hCoV-19/Ireland/G-NVRL-72IRL53006/2020\|EPI_ISL_578 | ···················· | ···················· | ···················· | ···················· | NNNNNNNNNNNNNNNNNNNN | NNNNNNNNNNNNNNNNNNNN | 1 |
| L | 5209 | hCoV-19/New Zealand/20VR3760/2020\|EPI_ISL_579507\|2 | ···················· | ···················· | ···················· | ···················· | NNNNNNNNNNNNNNNNNNNN | NNNNNNNNNNNNNNNNNNNN | 1 |
| L | 5210 | hCoV-19/New Zealand/20VR3761/2020\|EPI_ISL_579508\|2 | ···················· | ···················· | ···················· | ···················· | NNNNNNNNNNNNNNNNNNNN | NNNNNNNNNNNNNNNNNNNN | 1 |
| L | 5211 | hCoV-19/New Zealand/20VR3762/2020\|EPI_ISL_579509\|2 | ···················· | ···················· | ···················· | ···················· | NNNNNNNNNNNNNNNNNNNN | NNNNNNNNNNNNNNNNNNNN | 1 |
| L | 5212 | hCoV-19/New Zealand/20VR3763/2020\|EPI_ISL_579510\|2 | ···················· | ···················· | ···················· | ···················· | NNNNNNNNNNNNNNNNNNNN | NNNNNNNNNNNNNNNNNNNN | 1 |
| L | 5214 | hCoV-19/New Zealand/20VR3767/2020\|EPI_ISL_579513\|2 | ···················· | ···················· | ···················· | ···················· | NNNNNNNNNNNNNNNNNNNN | NNNNNNNNNNNNNNNNNNNN | 1 |
| L | 5340 | hCoV-19/Switzerland/BL-42217630/2020\|EPI_ISL_58182 | ···················· | ···················· | ···················· | ···················· | ·NNNNNNNN··········· | ···················· | 1 |
| L | 5358 | hCoV-19/Switzerland/BS-42220622/2020\|EPI_ISL_58185 | ···················· | ···················· | ···················· | ···················· | ·NNNNN·············· | ···················· | 1 |
| L | 5365 | hCoV-19/Switzerland/BL-65240191/2020\|EPI_ISL_58187 | ···················· | ···················· | ···················· | ···················· | ·NNNNNN············· | ···················· | 1 |
| L | 5366 | hCoV-19/Switzerland/BS-42341282/2020\|EPI_ISL_58187 | ···················· | ···················· | ···················· | ···················· | ·NNNNNNNNNNNNNNNNNNN | NNNNNNNNNNNNNNNNNNNN | 1 |
| L | 5367 | hCoV-19/Switzerland/BS-42343508/2020\|EPI_ISL_58188 | ···················· | ···················· | ···················· | ···················· | ·NNNNNNNNNNNN······· | ···················· | 1 |
| L | 5368 | hCoV-19/Switzerland/BS-42346247/2020\|EPI_ISL_58188 | ···················· | ···················· | ···················· | ···················· | NNNNNNNNNNNNNNNNNNNN | NNNNNNNNNNNNNNNNNNNN | 1 |
| L | 5369 | hCoV-19/Switzerland/BS-42302300/2020\|EPI_ISL_58189 | ···················· | ···················· | ···················· | ···················· | NNNNNNNNNNNNNNNNNNNN | NNNNNNNNNNNNNNNNNNNN | 1 |
| L | 5372 | hCoV-19/Switzerland/BS-42340959/2020\|EPI_ISL_58189 | ···················· | ···················· | ···················· | ···················· | NNNNNNNNNNNNNNNNNNNN | ···················· | 1 |
| L | 5373 | hCoV-19/Switzerland/BL-42340977/2020\|EPI_ISL_58189 | ···················· | ···················· | ···················· | ···················· | NNNNNNNNNNNNNNNNNNNN | NNNNNNNNNNNNNNNNNNNN | 1 |
| L | 5375 | hCoV-19/Switzerland/SG-42341324/2020\|EPI_ISL_58189 | ···················· | ···················· | ···················· | ···················· | NNNNNNNNNNNNNNNNNNNN | NNNN················ | 1 |
| L | 5465 | hCoV-19/England/NORW-EF1D7/2020\|EPI_ISL_573414\|202 | ···················· | ···················· | ···················· | ···················· | NNNNNNNNNNNNNNNNNNNN | NNNNNNNNNNNNNNNNNNNN | 1 |
| L | 5466 | hCoV-19/England/NORW-EF66F/2020\|EPI_ISL_573415\|202 | ···················· | ···················· | ···················· | ···················· | NNNNNNNNNNNNNNNNNNNN | NNNNNNNNNNNNNNNNNNNN | 1 |
| L | 5467 | hCoV-19/England/NORW-EF7C6/2020\|EPI_ISL_573416\|202 | ···················· | ···················· | ···················· | ···················· | NNNNNNNNNNNNNNNNNNNN | NNNNNNNNNNNNNNNNNNNN | 1 |
| L | 5468 | hCoV-19/England/NORW-EF993/2020\|EPI_ISL_573417\|202 | ···················· | ···················· | ···················· | ···················· | NNNNNNNNNNNNNNNNNNNN | NNNNNNNNNNNNNNNNNNNN | 1 |
| L | 5471 | hCoV-19/England/NOTT-1134B2/2020\|EPI_ISL_573420\|20 | ···················· | ···················· | ···················· | ···················· | NNNNNNNNNNNNNNNNNNNN | NNNNNNNNNNNNNNNNNNNN | 1 |
| L | 5472 | hCoV-19/England/NOTT-113704/2020\|EPI_ISL_573421\|20 | ···················· | ·A·················· | ···················· | ···················· | NNNNNNNNNNNNNNNNNNNN | NNNNNNNNNNNNNNNNNNNN | 1 |
| L | 5476 | hCoV-19/England/NOTT-11375F/2020\|EPI_ISL_573425\|20 | ···················· | NNNNNNNNNNNNNNNNNNNN | ···················· | ···················· | ···················· | ···················· | 1 |
| L | 5479 | hCoV-19/England/NOTT-11379B/2020\|EPI_ISL_573428\|20 | ···················· | NNNNNNNNNNNNNNNNNNNN | ···················· | ···················· | ···················· | ···················· | 1 |
| L | 5482 | hCoV-19/England/NOTT-1137D7/2020\|EPI_ISL_573431\|20 | ···················· | ···················· | ···················· | ···················· | NNNNNNNNNNNNNNNNNNNN | NNNNNNNNNNNNNNNNNNNN | 1 |
| L | 5498 | hCoV-19/England/NOTT-113977/2020\|EPI_ISL_573447\|20 | ···················· | ···················· | ···················· | ···················· | NNNNNNNNNNNNNNNNNNNN | NNNNNNNNNNNNNNNNNNNN | 1 |
| L | 5553 | hCoV-19/USA/MI-UM-10033815825/2020\|EPI_ISL_578130\| | ···················· | ···················· | ···················· | ···················· | NNNNNNNNNNNNNNNNNNNN | NNNNNNNNNNNNNNNNNNNN | 1 |
| L | 5558 | hCoV-19/USA/MI-UM-10033833216/2020\|EPI_ISL_578136\| | ···················· | ···················· | ···················· | ···················· | NNNNNNNNNNNNNNNNNNNN | ···················· | 1 |
| L | 5561 | hCoV-19/USA/MI-UM-10033851156/2020\|EPI_ISL_578139\| | ···················· | ···················· | ···················· | ···················· | NNNNNNNNNNNNNNNNNNNN | ···················· | 1 |
| L | 5563 | hCoV-19/USA/MI-UM-10033852420/2020\|EPI_ISL_578141\| | ···················· | ···················· | ···················· | ···················· | NNNNNNNNNNNNNNNNNNNN | ···················· | 1 |
| L | 5565 | hCoV-19/USA/MI-UM-10033866753/2020\|EPI_ISL_578143\| | ···················· | ···················· | ···················· | ···················· | NNNNNNNNNNNNNNNNNNNN | ···················· | 1 |
| L | 5566 | hCoV-19/USA/MI-UM-10033871564/2020\|EPI_ISL_578144\| | ···················· | ···················· | ···················· | ···················· | NNNNNNNNNNNNNNNNNNNN | NNNNNNNNNNNNNNNNNNNN | 1 |
| L | 5572 | hCoV-19/New Zealand/20VR3170/2020\|EPI_ISL_579460\|2 | ···················· | ···················· | ···················· | ···················· | NNNNNNNNNNNNNNNNNNNN | NNNNNNNNNNNNNNNNNNNN | 1 |
| L | 5579 | hCoV-19/New Zealand/20VR3178/2020\|EPI_ISL_579467\|2 | ···················· | ···················· | ···················· | ···················· | NNNNNNNNNNNNNNNNNNNN | NNNNNNNNNNNNNNNNNNNN | 1 |
| L | 5587 | hCoV-19/New Zealand/20VR3192/2020\|EPI_ISL_579479\|2 | ···················· | ···················· | ···················· | ···················· | NNNNNNNNNNNNNNNNNNNN | NNNNNNNNNNNNNNNNNNNN | 1 |
| L | 5615 | hCoV-19/Switzerland/BS-42351043/2020\|EPI_ISL_58190 | ···················· | ···················· | ···················· | ···················· | NNNNNNNNNNNNNNNNNNNN | NNNNNNNNNNNNNNNNNNNN | 1 |
| L | 5616 | hCoV-19/Switzerland/BS-65245511/2020\|EPI_ISL_58190 | ···················· | ···················· | ···················· | ···················· | NNNNNNNNNNNNNNNNNNNN | NNNNNNNNNNNNNNNNNNNN | 1 |
| L | 5617 | hCoV-19/Switzerland/BS-65246128/2020\|EPI_ISL_58190 | ···················· | ···················· | ···················· | ···················· | NNNNNNNNNNNNNNNNNNNN | NNNNNNNNNNNNNNNNNNNN | 1 |
| L | 5618 | hCoV-19/Switzerland/BL-65252178/2020\|EPI_ISL_58190 | ···················· | ···················· | ···················· | ···················· | NNNNNNNNNNNNNNNNNNNN | NNNNNNNNNNNNNNNNNNNN | 1 |
| L | 5619 | hCoV-19/Switzerland/BS-65253015/2020\|EPI_ISL_58190 | ···················· | ···················· | ···················· | ···················· | NNNNNNNNNNNNNNNNNNNN | NNNNNNNNNNNNNNNNNNNN | 1 |

```
L 5621 hCoV-19/Switzerland/BS-42361924/2020|EPI_ISL_58191 ···················· ···················· ···················· ···················· NNNNNNNNNNNNNNNNNNNN NNNNNNNNNNNNNNNNNNNN 1
L 5622 hCoV-19/Switzerland/BL-42176229/2020|EPI_ISL_58191 ···················· ···················· ···················· ···················· NNNNNNNNNNNNNNNNNNNN NNNNNNNNNNNNNNNNNNNN 1
L 5625 hCoV-19/Switzerland/AG-42243983/2020|EPI_ISL_58192 ···················· ···················· ···········G········ ···················· NNNNNNNNNNNNNNNNNNNN NNNNNNNNNNNNNNNNNNNN 1
L 5626 hCoV-19/Switzerland/BS-42248649/2020|EPI_ISL_58192 ···················· ···················· ···················· ···················· NNNNNNNNNNNNNNNNNNNN NNNNNNNNNNNNNNNNNNNN 1
L 5629 hCoV-19/Switzerland/BS-42256492/2020|EPI_ISL_58192 ···················· ···················· ···················· ···················· NNNNNNNNNNNNNNNNNNNN ···················· 1
L 5630 hCoV-19/Switzerland/BS-42260094/2020|EPI_ISL_58192 ···················· ···················· ···················· ···················· NNNNNNNNNNNNNNNNNNNN NNNNNNNNNNNNNNNNNNNN 1
L 5631 hCoV-19/Switzerland/BS-42276346/2020|EPI_ISL_58192 ···················· ···················· ···················· ···················· NNNNNNNNNNNNNNNNNNNN NNNNNNNNNNNNNNNNN··· 1
L 5632 hCoV-19/Switzerland/BL-42283924/2020|EPI_ISL_58192 ···················· ···················· ···················· ···················· NNNNNNNNNNNNNNNNNNNN ···················· 1
L 5664 hCoV-19/USA/MI-UM-10033797804/2020|EPI_ISL_578121| ···················· ···················· ···················· ···················· NNNNNNNNNNNNNNNNNNNN NNNNNNNNNNNNNNNNNNNN 1
L 5666 hCoV-19/USA/MI-UM-10033808341/2020|EPI_ISL_578123| ···················· ···················· ···················· ···················· NNNNNNNNNNNNNNNNNNNN ···················· 1
L 5675 hCoV-19/USA/MI-UM-10033901123/2020|EPI_ISL_578152| ···················· ···················· ···················· ···················· NNNNNNNNNNNNNNNNNNNN ···················· 1
L 5676 hCoV-19/USA/MI-UM-10033903914/2020|EPI_ISL_578153| ···················· ···················· ···················· ···················· ·······NNNNNNNNNNNNN ···················· 1
L 5677 hCoV-19/USA/MI-UM-10033906276/2020|EPI_ISL_578154| ···················· ···················· ···················· ···················· NNNNNNNNNNNNNNNNNNNN NNNNNNNNNNNNNNNNNNNN 1
L 5680 hCoV-19/USA/MI-UM-10033916721/2020|EPI_ISL_578157| ···················· ···················· ···················· ···················· NNNNNNNNNNNNNNNNNNNN ···················· 1
L 5681 hCoV-19/USA/MI-UM-10033920353/2020|EPI_ISL_578158| ···················· ···················· ···················· ···················· NNNNNNNNNNNNNNNNNNNN ···················· 1
L 5682 hCoV-19/USA/MI-UM-10033928982/2020|EPI_ISL_578159| ···················· ···················· ···················· ···················· NNNNNNNNNNNNNNNNNNNN NNNNNNNNNNNNNNNNNNNN 1
L 5688 hCoV-19/USA/MI-UM-10034006376/2020|EPI_ISL_578165| ···················· ···················· ···················· ···················· NNNNNNNNNNNNNNNNNNNN NNNNNNNNNNNNNNNNNNNN 1
L 5690 hCoV-19/USA/MI-UM-10034012043/2020|EPI_ISL_578167| ···················· ···················· ···················· ···················· NNNNNNNNNNNNNNNNNNNN NNNNNNNNNNNNNNNNNNNN 1
L 5692 hCoV-19/USA/MI-UM-10034056764/2020|EPI_ISL_578169| ···················· ···················· ···················· ···················· NNNNNNNNNNNNNNNNNNNN NNNNNNNNNNNNNNNNNNNN 1
L 5693 hCoV-19/USA/MI-UM-10034110078/2020|EPI_ISL_578170| ···················· ···················· ···················· ···················· NNNNNNNNNNNNNNNNNNNN ···················· 1
L 5695 hCoV-19/USA/MI-UM-10034148754/2020|EPI_ISL_578172| ···················· ···················· ···················· ···················· NNNNNNNNNNNNNNNNNNNN ···················· 1
L 5696 hCoV-19/USA/MI-UM-10034240230/2020|EPI_ISL_578173| ···················· ···················· ···················· ···················· NNNNNNNNNNNNNNNNNNNN ···················· 1
L 5708 hCoV-19/USA/MI-UM-10033991503/2020|EPI_ISL_578185| ···················· ···················· ···················· ···················· NNNNNNNNNNNNNNNNNNNN ···················· 1
L 5724 hCoV-19/New Zealand/20VR3044/2020|EPI_ISL_579402|2 ···················· ···················· ···················· ···················· NNNNNNNNNNNNNNNNNNNN NNNNNNNNNNNNNNNNNNNN 1
L 5725 hCoV-19/New Zealand/20VR3060/2020|EPI_ISL_579403|2 ···················· ···················· ···················· ···················· NNNNNNNNNNNNNNNNNNNN NNNNNNNNNNNNNNNNNNNN 1
L 5726 hCoV-19/New Zealand/20VR3063/2020|EPI_ISL_579404|2 ···················· ···················· ···················· ···················· NNNNNNNNNNNNNNNNNNNN NNNNNNNNNNNNNNNNNNNN 1
L 5728 hCoV-19/New Zealand/20VR3066/2020|EPI_ISL_579406|2 ···················· ···················· ···················· ···················· NNNNNNNNNNNNNNNNNNNN NNNNNNNNNNNNNNNNNNNN 1
L 5730 hCoV-19/New Zealand/20VR3070/2020|EPI_ISL_579408|2 ···················· ···················· ···················· ···················· NNNNNNNNNNNNNNNNNNNN NNNNNNNNNNNNNNNNNNNN 1
L 5731 hCoV-19/New Zealand/20VR3071/2020|EPI_ISL_579409|2 ···················· ···················· ···················· ···················· NNNNNNNNNNNNNNNNNNNN NNNNNNNNNNNNNNNNNNNN 1
L 5732 hCoV-19/New Zealand/20VR3073/2020|EPI_ISL_579410|2 ···················· ···················· ···················· ···················· NNNNNNNNNNNNNNNNNNNN NNNNNNNNNNNNNNNNNNNN 1
L 5733 hCoV-19/New Zealand/20VR3074/2020|EPI_ISL_579411|2 ···················· ···················· ···················· ···················· NNNNNNNNNNNNNNNNNNNN NNNNNNNNNNNNNNNNNNNN 1
L 5735 hCoV-19/New Zealand/20VR3084/2020|EPI_ISL_579414|2 ···················· ···················· ···················· ···················· NNNNNNNNNNNNNNNNNNNN NNNNNNNNNNNNNNNNNNNN 1
L 5737 hCoV-19/New Zealand/20VR3089/2020|EPI_ISL_579416|2 ···················· ···················· ···················· ···················· NNNNNNNNNNNNNNNNNNNN NNNNNNNNNNNNNNNNNNNN 1
L 5741 hCoV-19/New Zealand/20VR3099/2020|EPI_ISL_579420|2 ···················· ···················· ···················· ···················· NNNNNNNNNNNNNNNNNNNN NNNNNNNNNNNNNNNNNNNN 1
L 5743 hCoV-19/New Zealand/20VR3112/2020|EPI_ISL_579422|2 ···················· ···················· ···················· ···················· NNNNNNNNNNNNNNNNNNNN NNNNNNNNNNNNNNNNNNNN 1
L 5748 hCoV-19/New Zealand/20VR3135/2020|EPI_ISL_579427|2 ···················· ···················· ···················· ···················· NNNNNNNNNNNNNNNNNNNN NNNNNNNNNNNNNNNNNNNN 1
L 5749 hCoV-19/New Zealand/20VR3137/2020|EPI_ISL_579429|2 ···················· ···················· ···················· ···················· NNNNNNNNNNNNNNNNNNNN NNNNNNNNNNNNNNNNNNNN 1
L 5751 hCoV-19/New Zealand/20VR3139/2020|EPI_ISL_579431|2 ···················· ···················· ···················· ···················· NNNNNNNNNNNNNNNNNNNN NNNNNNNNNNNNNNNNNNNN 1
L 5752 hCoV-19/New Zealand/20VR3140/2020|EPI_ISL_579432|2 ···················· ···A················ ···················· ···················· NNNNNNNNNNNNNNNNNNNN NNNNNNNNNNNNNNNNNNNN 1
L 5753 hCoV-19/New Zealand/20VR3141/2020|EPI_ISL_579433|2 ···················· ···A················ ···················· ···················· NNNNNNNNNNNNNNNNNNNN NNNNNNNNNNNNNNNNNNNN 1
L 5754 hCoV-19/New Zealand/20VR3142/2020|EPI_ISL_579434|2 ···················· ·A·················· ···················· ···················· NNNNNNNNNNNNNNNNNNNN NNNNNNNNNNNNNNNNNNNN 1
L 5756 hCoV-19/New Zealand/20VR3144/2020|EPI_ISL_579436|2 ···················· ···················· ···················· ···················· NNNNNNNNNNNNNNNNNNNN NNNNNNNNNNNNNNNNNNNN 1
L 5757 hCoV-19/New Zealand/20VR3145/2020|EPI_ISL_579437|2 ···················· ···················· ···················· ···················· NNNNNNNNNNNNNNNNNNNN NNNNNNNNNNNNNNNNNNNN 1
L 5758 hCoV-19/New Zealand/20VR3162/2020|EPI_ISL_579452|2 ···················· ···················· ···················· ···················· NNNNNNNNNNNNNNNNNNNN NNNNNNNNNNNNNNNNNNNN 1
L 5760 hCoV-19/New Zealand/20VR3165/2020|EPI_ISL_579455|2 ···················· ···················· ···················· ···················· NNNNNNNNNNNNNNNNNNNN NNNNNNNNNNNNNNNNNNNN 1
L 5762 hCoV-19/New Zealand/20VR3167/2020|EPI_ISL_579457|2 ···················· ···················· ···················· ···················· NNNNNNNNNNNNNNNNNNNN NNNNNNNNNNNNNNNNNNNN 1
L 5768 hCoV-19/New Zealand/20VR3197/2020|EPI_ISL_579483|2 ···················· ···················· ···················· ···················· NNNNNNNNNNNNNNNNNNNN NNNNNNNNNNNNNNNNNNNN 1
L 5769 hCoV-19/New Zealand/20VR3198/2020|EPI_ISL_579484|2 ···················· ···················· ···················· ···················· NNNNNNNNNNNNNNNNNNNN NNNNNNNNNNNNNNNNNNNN 1
L 5771 hCoV-19/New Zealand/20VR3201/2020|EPI_ISL_579486|2 ···················· ···················· ···················· ···················· NNNNNNNNNNNNNNNNNNNN NNNNNNNNNNNNNNNNNNNN 1
L 5774 hCoV-19/New Zealand/20VR3204/2020|EPI_ISL_579489|2 ···················· ···················· ···················· ···················· NNNNNNNNNNNNNNNNNNNN NNNNNNNNNNNNNNNNNNNN 1
L 5775 hCoV-19/New Zealand/20VR3206/2020|EPI_ISL_579491|2 ···················· ···················· ···················· ···················· NNNNNNNNNNNNNNNNNNNN NNNNNNNNNNNNNNNNNNNN 1
L 5776 hCoV-19/New Zealand/20VR3207/2020|EPI_ISL_579492|2 ···················· ···················· ···················· ···················· NNNNNNNNNNNNNNNNNNNN NNNNNNNNNNNNNNNNNNNN 1
L 5861 hCoV-19/Switzerland/BS-42206450/2020|EPI_ISL_58175 ···················· ···················· ···················· ···················· NNNN················ ···················· 1
L 5862 hCoV-19/Switzerland/BS-42206747/2020|EPI_ISL_58175 ···················· ···················· ···················· ···················· NNNNNNNNNNNNNNNNNNNN NNNNNNNNNNNNNNNNNNNN 1
L 5864 hCoV-19/Switzerland/BS-42207789/2020|EPI_ISL_58175 ···················· ···················· ···················· ···················· NNNNNNNNNNNNNNNNNNNN NNNNNNNNNNNNNNNNNNNN 1
L 5866 hCoV-19/Switzerland/BS-42207989/2020|EPI_ISL_58175 ···················· ···················· ···················· ···················· NNNNNNNNNNNNNNNNNNNN NNNNNNNNNNNNNNNNNNNN 1
L 5869 hCoV-19/Switzerland/BS-42208210/2020|EPI_ISL_58176 ···················· ···················· ···················· ···················· NNNNNNNNNNNNNNNNNNNN NNNNNNNNNNNNNNNNNNNN 1
L 5870 hCoV-19/Switzerland/BL-42208262/2020|EPI_ISL_58176 ···················· ···················· ···················· ···················· NNNNNNNNNNNNNNNNNNNN NNNNNNNNNNNNNNNNNNNN 1
L 5873 hCoV-19/Switzerland/BL-42208623/2020|EPI_ISL_58177 ···················· ···················· ···················· ···················· NNNNNNNNNNNNNNNNNNNN NNNNNNNNNNNNNNNNNNNN 1
L 5875 hCoV-19/Switzerland/BS-42209439/2020|EPI_ISL_58177 ···················· ···················· ···················· ···················· NNNNNNNNNNNNNNNNNNNN NNNNNNNNNNNNNNNNNNNN 1
L 5876 hCoV-19/Switzerland/BS-42209452/2020|EPI_ISL_58177 ···················· ···················· ···················· ···················· NNNNNNNNNNNNNNNNNNNN NNNNNNNNNNNNNNNNNNNN 1
L 5878 hCoV-19/Switzerland/BS-42209528/2020|EPI_ISL_58178 ···················· ···················· ···················· ···················· NNNNNNNNNNNNNNNNNNNN NNNNNNNNNNNNNNNNNNNN 1
L 5963 hCoV-19/England/MILK-9C736D/2020|EPI_ISL_573300|20 ···················· ···················· ···················· ···················· NNNNNNNNNNNNNNNNNNNN NNNNNNNNNNNNNNNNNNNN 1
L 5981 hCoV-19/England/NORT-2A0C66/2020|EPI_ISL_573362|20 ···················· ···················· ···················· ···················· NNNNNNNNNNNNNNNNNNNN NNNNNNNNNNNNNNNNNNNN 1
L 5983 hCoV-19/England/NORT-2A0CFD/2020|EPI_ISL_573364|20 ···················· ···················· ···················· ···················· NNNNNNNNNNNNNNNNNNNN NNNNNNNNNNNNNNNNNNNN 1
L 5985 hCoV-19/England/NORT-2A0D36/2020|EPI_ISL_573366|20 ···················· ···················· ···················· ···················· NNNNNNNNNNNNNNNNNNNN NNNNNNNNNNNNNNNNNNNN 1
L 5993 hCoV-19/England/NORT-2A43E5/2020|EPI_ISL_573374|20 ···················· ···················· ···················· ···················· NNNNNNNNNNNNNNNNNNNN NNNNNNNNNNNNNNNNNNNN 1
L 6087 hCoV-19/USA/MI-UM-10033785306/2020|EPI_ISL_578111| ···················· ···················· ···················· ···················· NNNNNNNNNNNNNNNNNNNN ···················· 1
L 6089 hCoV-19/USA/MI-UM-10033789519/2020|EPI_ISL_578113| ···················· ···················· ···················· ···················· NNNNNNNNNNNNNNNNNNNN ···················· 1
L 6093 hCoV-19/USA/MI-UM-10033792777/2020|EPI_ISL_578117| ···················· ···················· ···················· ···················· NNNNNNNNNNNNNNNNNNNN ···················· 1
L 6095 hCoV-19/USA/MI-UM-10033793415/2020|EPI_ISL_578119| ···················· ···················· ···················· ···················· NNNNNNNNNNNNNNNNNNNN NNNNNNNNNNNNNNNNNNNN 1
L 6096 hCoV-19/USA/MI-UM-10033797461/2020|EPI_ISL_578120| ···················· ···················· ···················· ···················· NNNNNNNNNNNNNNNNNNNN NNNNNNNNNNNNNNNNNNNN 1
L 6098 hCoV-19/New Zealand/20VR3146/2020|EPI_ISL_579438|2 ···················· ···················· ···················· ···················· NNNNNNNNNNNNNNNNNNNN NNNNNNNNNNNNNNNNNNNN 1
L 6099 hCoV-19/New Zealand/20VR3147/2020|EPI_ISL_579439|2 ···················· ···················· ···················· ···················· NNNNNNNNNNNNNNNNNNNN NNNNNNNNNNNNNNNNNNNN 1
L 6102 hCoV-19/New Zealand/20VR3151/2020|EPI_ISL_579442|2 ···················· ···················· ···················· ···················· NNNNNNNNNNNNNNNNNNNN NNNNNNNNNNNNNNNNNNNN 1
L 6103 hCoV-19/New Zealand/20VR3152/2020|EPI_ISL_579443|2 ···················· ···················· ···················· ···················· NNNNNNNNNNNNNNNNNNNN NNNNNNNNNNNNNNNNNNNN 1
L 6106 hCoV-19/New Zealand/20VR3156/2020|EPI_ISL_579446|2 ···················· ···················· ···················· ···················· NNNNNNNNNNNNNNNNNNNN NNNNNNNNNNNNNNNNNNNN 1
L 6107 hCoV-19/New Zealand/20VR3157/2020|EPI_ISL_579447|2 ···················· ···················· ···················· ···················· NNNNNNNNNNNNNNNNNNNN NNNNNNNNNNNNNNNNNNNN 1
L 6109 hCoV-19/New Zealand/20VR3160/2020|EPI_ISL_579450|2 ···················· ···················· ···················· ···················· NNNNNNNNNNNNNNNNNNNN NNNNNNNNNNNNNNNNNNNN 1
L 6110 hCoV-19/New Zealand/20VR3161/2020|EPI_ISL_579451|2 ···················· ···················· ···················· ···················· NNNNNNNNNNNNNNNNNNNN NNNNNNNNNNNNNNNNNNNN 1
```

| | | | | | | | | |
|---|---|---|---|---|---|---|---|---|
| L | 6129 hCoV-19/Switzerland/BS-42204906/2020\|EPI_ISL_58173 | ···················· | ···················· | ···················· | ···················· | NNNNNNNNNNNNNNNNNNNN | NNNN················ | 1 |
| L | 6131 hCoV-19/Switzerland/BS-42205257/2020\|EPI_ISL_58174 | ···················· | ···················· | ···················· | ···················· | ·NN················· | NNNNNNNNN··········· | 1 |
| L | 6137 hCoV-19/Switzerland/BS-42206374/2020\|EPI_ISL_58174 | ···················· | ···················· | ···················· | ···················· | ·NNNNN·············· | ···················· | 1 |
| L | 6140 hCoV-19/Switzerland/ZH-42209655/2020\|EPI_ISL_58180 | ···················· | ···················· | ···················· | ···················· | NNNNNNNNNNNNNNNNNNNN | ···················· | 1 |
| L | 6141 hCoV-19/Switzerland/BS-42210842/2020\|EPI_ISL_58180 | ···················· | ···················· | ···················· | ···················· | NNNNNNNNNNNNNNNNNNNN | NNNN················ | 1 |
| L | 6144 hCoV-19/Switzerland/BS-42210930/2020\|EPI_ISL_58180 | ···················· | ···················· | ···················· | ···················· | NNNNNNNNNNNNNNNNNNNN | NNNNNNNNNNNNNNNNNNNN | 1 |
| L | 6182 hCoV-19/England/NORT-293CFD/2020\|EPI_ISL_573324\|20 | ···················· | ···················· | ···················· | ···················· | NNNNNNNNNNNNNNNNNNNN | NNNNNNNNNNNNNNNNNNNN | 1 |
| L | 6190 hCoV-19/England/NORT-293F21/2020\|EPI_ISL_573332\|20 | ···················· | ···················· | ···················· | ···················· | NNNNNNNNNNNNNNNNNNNN | NNNNNNNNNNNNNNNNNNNN | 1 |
| L | 6195 hCoV-19/England/NORT-294157/2020\|EPI_ISL_573337\|20 | ···················· | ···················· | ···················· | ···················· | NNNNNNNNNNNNNNNNNNNN | NNNNNNNNNNNNNNNNNNNN | 1 |
| L | 6196 hCoV-19/England/NORT-294166/2020\|EPI_ISL_573338\|20 | ···················· | ···················· | ···················· | ···················· | NNNNNNNNNNNNNNNNNNNN | NNNNNNNNNNNNNNNNNNNN | 1 |
| L | 6305 hCoV-19/New Zealand/20VR2722/2020\|EPI_ISL_579340\|2 | ···················· | ···················· | ···················· | ···················· | NNNNNNNNNNNNNNNNNNNN | NNNNNNNNNNNNNNNNNNNN | 1 |
| L | 6307 hCoV-19/New Zealand/20VR2727/2020\|EPI_ISL_579344\|2 | ···················· | ···················· | ···················· | ···················· | NNNNNNNNNNNNNNNNNNNN | NNNNNNNNNNNNNNNNNNNN | 1 |
| L | 6308 hCoV-19/New Zealand/20VR2730/2020\|EPI_ISL_579345\|2 | ···················· | ···················· | ···················· | ···················· | NNNNNNNNNNNNNNNNNNNN | NNNNNNNNNNNNNNNNNNNN | 1 |
| L | 6309 hCoV-19/New Zealand/20VR2731/2020\|EPI_ISL_579346\|2 | ···················· | ···················· | ···················· | ···················· | NNNNNNNNNNNNNNNNNNNN | NNNNNNNNNNNNNNNNNNNN | 1 |
| L | 6312 hCoV-19/New Zealand/20VR2920/2020\|EPI_ISL_579351\|2 | ···················· | ···················· | ···················· | ···················· | NNNNNNNNNNNNNNNNNNNN | NNNNNNNNNNNNNNNNNNNN | 1 |
| L | 6313 hCoV-19/New Zealand/20VR2923/2020\|EPI_ISL_579352\|2 | ···················· | ···················· | ···················· | ···················· | NNNNNNNNNNNNNNNNNNNN | NNNNNNNNNNNNNNNNNNNN | 1 |
| L | 6314 hCoV-19/New Zealand/20VR2925/2020\|EPI_ISL_579354\|2 | ···················· | ···················· | ···················· | ···················· | NNNNNNNNNNNNNNNNNNNN | NNNNNNNNNNNNNNNNNNNN | 1 |
| L | 6317 hCoV-19/New Zealand/20VR2935/2020\|EPI_ISL_579359\|2 | ···················· | ···················· | ···················· | ···················· | NNNNNNNNNNNNNNNNNNNN | NNNNNNNNNNNNNNNNNNNN | 1 |
| L | 6321 hCoV-19/New Zealand/20VR2956/2020\|EPI_ISL_579375\|2 | ···················· | ···················· | ···················· | ···················· | NNNNNNNNNNNNNNNNNNNN | NNNNNNNNNNNNNNNNNNNN | 1 |
| L | 6322 hCoV-19/New Zealand/20VR2957/2020\|EPI_ISL_579376\|2 | ···················· | ···················· | ···················· | ···················· | NNNNNNNNNNNNNNNNNNNN | NNNNNNNNNNNNNNNNNNNN | 1 |
| L | 6359 hCoV-19/Belgium/ULG-10488/2020\|EPI_ISL_581651\|2020 | ···················· | ···················· | ···················· | ···················· | NNNNNNNNNNNNNNNNNNNN | NNNNNNNNNNNNNNNNNNNN | 1 |
| L | 6360 hCoV-19/Belgium/ULG-10489/2020\|EPI_ISL_581652\|2020 | ···················· | ···················· | ···················· | ···················· | NNNNNNNNNNNNNNNNNNNN | NNNNNNNNNNNNNNNNNNNN | 1 |
| L | 6364 hCoV-19/Belgium/ULG-10494/2020\|EPI_ISL_581656\|2020 | ···················· | ···················· | ···················· | ···················· | NNNNNNNNNNNNNNNNNNNN | NNNNNNNNNNNNNNNNNNNN | 1 |
| L | 6365 hCoV-19/Belgium/ULG-10495/2020\|EPI_ISL_581657\|2020 | ···················· | ···················· | ···················· | ···················· | NNNNNNNNNNNNNNNNNNNN | NNNNNNNNNNNNNNNNNNNN | 1 |
| L | 6366 hCoV-19/Belgium/ULG-10496/2020\|EPI_ISL_581658\|2020 | ···················· | ···················· | ···················· | ···················· | NNNNNNNNNNNNNNNNNNNN | NNNNNNNNNNNNNNNNNNNN | 1 |
| L | 6368 hCoV-19/Belgium/ULG-10499/2020\|EPI_ISL_581661\|2020 | ···················· | ···················· | ···················· | ···················· | NNNNNNNNNNNNNNNNNNNN | NNNNNNNNNNNNNNNNNNNN | 1 |
| L | 6369 hCoV-19/Belgium/ULG-10500/2020\|EPI_ISL_581662\|2020 | ···················· | ···················· | ···················· | ···················· | NNNNNNNNNNNNNNNNNNNN | NNNNNNNNNNNNNNNNNNNN | 1 |
| L | 6371 hCoV-19/Belgium/ULG-10502/2020\|EPI_ISL_581664\|2020 | ···················· | ···················· | ···················· | ···················· | NNNNNNNNNNNNNNNNNNNN | NNNNNNNNNNNNNNNNNNNN | 1 |
| L | 6372 hCoV-19/Belgium/ULG-10503/2020\|EPI_ISL_581665\|2020 | ···················· | ···················· | ···················· | ···················· | NNNNNNNNNNNNNNNNNNNN | NNNNNNNNNNNNNNNNNNNN | 1 |
| L | 6373 hCoV-19/Belgium/ULG-10504/2020\|EPI_ISL_581666\|2020 | ···················· | ···················· | ···················· | ···················· | NNNNNNNNNNNNNNNNNNNN | NNNNNNNNNNNNNNNNNNNN | 1 |
| L | 6376 hCoV-19/Switzerland/BS-42216451/2020\|EPI_ISL_58167 | ···················· | ···················· | ···················· | ···················· | NNNNNNNNNNNNNNNNNNNN | NNNNNNNNNNNNNNNNNNNN | 1 |
| L | 6377 hCoV-19/Switzerland/BS-42216457/2020\|EPI_ISL_58167 | ···················· | ···················· | ···················· | ···················· | NNNNNNNNNNNNNNNNNNNN | NNNNNNNNNNNNNNNNNNNN | 1 |
| L | 6447 hCoV-19/New Zealand/20VR2572/2020\|EPI_ISL_579303\|2 | ···················· | ···················· | ···················· | ···················· | NNNNNNNNNNNNNNNNNNNN | NNNNNNNNNNNNNNNNNNNN | 1 |
| L | 6450 hCoV-19/New Zealand/20VR2575/2020\|EPI_ISL_579306\|2 | ···················· | ···················· | ···················· | ···················· | NNNNNNNNNNNNNNNNNNNN | NNNNNNNNNNNNNNNNNNNN | 1 |
| L | 6452 hCoV-19/New Zealand/20VR2577/2020\|EPI_ISL_579308\|2 | ···················· | ···················· | ···················· | ···················· | NNNNNNNNNNNNNNNNNNNN | NNNNNNNNNNNNNNNNNNNN | 1 |
| L | 6453 hCoV-19/New Zealand/20VR2578/2020\|EPI_ISL_579309\|2 | ···················· | ···················· | ···················· | ···················· | NNNNNNNNNNNNNNNNNNNN | NNNNNNNNNNNNNNNNNNNN | 1 |
| L | 6457 hCoV-19/New Zealand/20VR2647/2020\|EPI_ISL_579315\|2 | ···················· | ···················· | ···················· | ···················· | NNNNNNNNNNNNNNNNNNNN | NNNNNNNNNNNNNNNNNNNN | 1 |
| L | 6460 hCoV-19/New Zealand/20VR2714/2020\|EPI_ISL_579334\|2 | ···················· | ···················· | ···················· | ···················· | NNNNNNNNNNNNNNNNNNNN | NNNNNNNNNNNNNNNNNNNN | 1 |
| L | 6461 hCoV-19/New Zealand/20VR2716/2020\|EPI_ISL_579335\|2 | ···················· | ···················· | ···················· | ···················· | NNNNNNNNNNNNNNNNNNNN | NNNNNNNNNNNNNNNNNNNN | 1 |
| L | 6462 hCoV-19/New Zealand/20VR2719/2020\|EPI_ISL_579337\|2 | ···················· | ···················· | ···················· | ···················· | NNNNNNNNNNNNNNNNNNNN | NNNNNNNNNNNNNNNNNNNN | 1 |
| L | 6463 hCoV-19/New Zealand/20VR2720/2020\|EPI_ISL_579338\|2 | ···················· | ···················· | ···················· | ···················· | NNNNNNNNNNNNNNNNNNNN | NNNNNNNNNNNNNNNNNNNN | 1 |
| L | 6464 hCoV-19/New Zealand/20VR2936/2020\|EPI_ISL_579360\|2 | ···················· | ···················· | ···················· | ···················· | NNNNNNNNNNNNNNNNNNNN | NNNNNNNNNNNNNNNNNNNN | 1 |
| L | 6465 hCoV-19/New Zealand/20VR2937/2020\|EPI_ISL_579361\|2 | ···················· | ···················· | ···················· | ···················· | NNNNNNNNNNNNNNNNNNNN | NNNNNNNNNNNNNNNNNNNN | 1 |
| L | 6466 hCoV-19/New Zealand/20VR2940/2020\|EPI_ISL_579362\|2 | ···················· | ···················· | ···················· | ···················· | NNNNNNNNNNNNNNNNNNNN | NNNNNNNNNNNNNNNNNNNN | 1 |
| L | 6468 hCoV-19/New Zealand/20VR2946/2020\|EPI_ISL_579366\|2 | ···················· | ···················· | ···················· | ···················· | NNNNNNNNNNNNNNNNNNNN | NNNNNNNNNNNNNNNNNNNN | 1 |
| L | 6469 hCoV-19/New Zealand/20VR2947/2020\|EPI_ISL_579367\|2 | ···················· | ···················· | ···················· | ···················· | NNNNNNNNNNNNNNNNNNNN | NNNNNNNNNNNNNNNNNNNN | 1 |
| L | 6471 hCoV-19/New Zealand/20VR3009/2020\|EPI_ISL_579381\|2 | ···················· | ···················· | ···················· | ···················· | NNNNNNNNNNNNNNNNNNNN | NNNNNNNNNNNNNNNNNNNN | 1 |
| L | 6474 hCoV-19/New Zealand/20VR3012/2020\|EPI_ISL_579384\|2 | ···················· | ···················· | ···················· | ···················· | NNNNNNNNNNNNNNNNNNNN | NNNNNNNNNNNNNNNNNNNN | 1 |
| L | 6476 hCoV-19/New Zealand/20VR3017/2020\|EPI_ISL_579387\|2 | ···················· | ···················· | ···················· | ···················· | NNNNNNNNNNNNNNNNNNNN | NNNNNNNNNNNNNNNNNNNN | 1 |
| L | 6480 hCoV-19/New Zealand/20VR3025/2020\|EPI_ISL_579391\|2 | ···················· | ···················· | ···················· | ···················· | NNNNNNNNNNNNNNNNNNNN | NNNNNNNNNNNNNNNNNNNN | 1 |
| L | 6482 hCoV-19/New Zealand/20VR3030/2020\|EPI_ISL_579394\|2 | ···················· | ···················· | ···················· | ···················· | NNNNNNNNNNNNNNNNNNNN | NNNNNNNNNNNNNNNNNNNN | 1 |
| L | 6483 hCoV-19/New Zealand/20VR3031/2020\|EPI_ISL_579395\|2 | ···················· | ···················· | ···················· | ···················· | NNNNNNNNNNNNNNNNNNNN | NNNNNNNNNNNNNNNNNNNN | 1 |
| L | 6484 hCoV-19/New Zealand/20VR3032/2020\|EPI_ISL_579396\|2 | ···················· | ···················· | ···················· | ···················· | NNNNNNNNNNNNNNNNNNNN | NNNNNNNNNNNNNNNNNNNN | 1 |
| L | 6485 hCoV-19/New Zealand/20VR3033/2020\|EPI_ISL_579397\|2 | ···················· | ···················· | ···················· | ···················· | NNNNNNNNNNNNNNNNNNNN | NNNNNNNNNNNNNNNNNNNN | 1 |
| L | 6569 hCoV-19/Belgium/ULG-10465/2020\|EPI_ISL_581636\|2020 | ···················· | ···················· | ···················· | ···················· | NNNNNNNNNNNNNNNNNNNN | NNNNNNNNNNNNNNNNNNNN | 1 |
| L | 6570 hCoV-19/Belgium/ULG-10467/2020\|EPI_ISL_581637\|2020 | ···················· | ···················· | ···················· | ···················· | NNNNNNNNNNNNNNNNNNNN | NNNNNNNNNNNNNNNNNNNN | 1 |
| L | 6571 hCoV-19/Belgium/ULG-10468/2020\|EPI_ISL_581638\|2020 | ···················· | ···················· | ···················· | ···················· | NNNNNNNNNNNNNNNNNNNN | NNNNNNNNNNNNNNNNNNNN | 1 |
| L | 6573 hCoV-19/Belgium/ULG-10470/2020\|EPI_ISL_581640\|2020 | ···················· | ···················· | ···················· | ···················· | NNNNNNNNNNNNNNNNNNNN | NNNNNNNNNNNNNNNNNNNN | 1 |
| L | 6574 hCoV-19/Belgium/ULG-10471/2020\|EPI_ISL_581641\|2020 | ···················· | ···················· | ···················· | ···················· | NNNNNNNNNNNNNNNNNNNN | NNNNNNNNNNNNNNNNNNNN | 1 |
| L | 6575 hCoV-19/Belgium/ULG-10473/2020\|EPI_ISL_581642\|2020 | ···················· | ·A·················· | ···················· | ···················· | NNNNNNNNNNNNNNNNNNNN | NNNNNNNNNNNNNNNNNNNN | 1 |
| L | 6576 hCoV-19/Belgium/ULG-10474/2020\|EPI_ISL_581643\|2020 | ···················· | ···················· | ···················· | ···················· | NNNNNNNNNNNNNNNNNNNN | NNNNNNNNNNNNNNNNNNNN | 1 |
| L | 6578 hCoV-19/Belgium/ULG-10476/2020\|EPI_ISL_581645\|2020 | ···················· | ···················· | ···················· | ···················· | NNNNNNNNNNNNNNNNNNNN | NNNNNNNNNNNNNNNNNNNN | 1 |
| L | 6579 hCoV-19/Belgium/ULG-10478/2020\|EPI_ISL_581646\|2020 | ···················· | ···················· | ···················· | ···················· | NNNNNNNNNNNNNNNNNNNN | NNNNNNNNNNNNNNNNNNNN | 1 |
| L | 6580 hCoV-19/Belgium/ULG-10480/2020\|EPI_ISL_581647\|2020 | ···················· | ···A················ | ···················· | ···················· | NNNNNNNNNNNNNNNNNNNN | NNNNNNNNNNNNNNNNNNNN | 1 |
| L | 6582 hCoV-19/Belgium/ULG-10484/2020\|EPI_ISL_581649\|2020 | ···················· | ·A·················· | ···················· | ···················· | NNNNNNNNNNNNNNNNNNNN | NNNNNNNNNNNNNNNNNNNN | 1 |
| L | 6635 hCoV-19/USA/CA-CZB-5760/2020\|EPI_ISL_582964\|2020-0 | ···················· | ···················· | ···················· | ···················· | ··NNNNNNNNNNNNNNNNNN | ···················· | 1 |
| L | 6644 hCoV-19/USA/CA-CZB-5681/2020\|EPI_ISL_582973\|2020-0 | ···················· | ···················· | ···················· | ···················· | NNNNNNNNNNNNNNNNNNNN | NNNNNNNNNNNNNNNNNNNN | 1 |
| L | 6684 hCoV-19/England/MILK-9A7F50/2020\|EPI_ISL_573248\|20 | ···················· | ···················· | ···················· | ···················· | NNNNNNNNNNNNNNNNNNNN | NNNNNNNNNNNNNNNNNNNN | 1 |
| L | 6686 hCoV-19/England/MILK-9A7FBA/2020\|EPI_ISL_573250\|20 | ···················· | ···················· | ···················· | ···················· | NNNNNNNNNNNNNNNNNNNN | NNNNNNNNNNNNNNNNNNNN | 1 |
| L | 6705 hCoV-19/England/MILK-9C6F6A/2020\|EPI_ISL_573269\|20 | ···················· | ···················· | ···················· | ···················· | NNNNNNNNNNNNNNNNNNNN | NNNNNNNNNNNNNNNNNNNN | 1 |
| L | 6711 hCoV-19/England/MILK-9C7075/2020\|EPI_ISL_573275\|20 | ···················· | ···················· | ···················· | ···················· | NNNNNNNNNNNNNNNNNNNN | NNNNNNNNNNNNNNNNNNNN | 1 |
| L | 6712 hCoV-19/England/MILK-9C7084/2020\|EPI_ISL_573276\|20 | ···················· | ···················· | ···················· | ···················· | NNNNNNNNNNNNNNNNNNNN | NNNNNNNNNNNNNNNNNNNN | 1 |
| L | 6717 hCoV-19/England/MILK-9C70EE/2020\|EPI_ISL_573281\|20 | ···················· | ···················· | ···················· | ···················· | NNNNNNNNNNNNNNNNNNNN | NNNNNNNNNNNNNNNNNNNN | 1 |
| L | 6718 hCoV-19/England/MILK-9C7136/2020\|EPI_ISL_573282\|20 | ···················· | ···················· | ···················· | ···················· | NNNNNNNNNNNNNNNNNNNN | NNNNNNNNNNNNNNNNNNNN | 1 |
| L | 6721 hCoV-19/England/MILK-9C7190/2020\|EPI_ISL_573285\|20 | ···················· | ···················· | ···················· | ···················· | NNNNNNNNNNNNNNNNNNNN | NNNNNNNNNNNNNNNNNNNN | 1 |
| L | 6730 hCoV-19/England/MILK-9C72E8/2020\|EPI_ISL_573294\|20 | ···················· | ·A·················· | ···················· | ···················· | NNNNNNNNNNNNNNNNNNNN | NNNNNNNNNNNNNNNNNNNN | 1 |
| L | 6873 hCoV-19/USA/MI-UM-10033851236/2020\|EPI_ISL_578091\| | ···················· | ···················· | ···················· | ···················· | NNNNNNNNNNNNNNNNNNNN | ···················· | 1 |
| L | 6874 hCoV-19/USA/MI-UM-10033798803/2020\|EPI_ISL_578093\| | ···················· | ···················· | ···················· | ···················· | NNNNNNNNNNNNNNNNNNNN | NNNNNNNNNNNNNNNNNNNN | 1 |
| L | 6881 hCoV-19/New Zealand/20VR2652/2020\|EPI_ISL_579320\|2 | ···················· | ···················· | ···················· | ···················· | NNNNNNNNNNNNNNNNNNNN | NNNNNNNNNNNNNNNNNNNN | 1 |
| L | 6882 hCoV-19/New Zealand/20VR2653/2020\|EPI_ISL_579321\|2 | ···················· | ···················· | ···················· | ···················· | NNNNNNNNNNNNNNNNNNNN | NNNNNNNNNNNNNNNNNNNN | 1 |

| | | | | | | | | |
|---|---|---|---|---|---|---|---|---|
| L | 6883 hCoV-19/New Zealand/20VR2654/2020\|EPI_ISL_579322\|2 | ···················· | ···················· | ···················· | ···················· | NNNNNNNNNNNNNNNNNNNN | NNNNNNNNNNNNNNNNNNNN | 1 |
| L | 6884 hCoV-19/New Zealand/20VR2656/2020\|EPI_ISL_579323\|2 | ···················· | ···················· | ···················· | ···················· | NNNNNNNNNNNNNNNNNNNN | NNNNNNNNNNNNNNNNNNNN | 1 |
| L | 6887 hCoV-19/Belgium/ULG-10435/2020\|EPI_ISL_581613\|2020 | ···················· | ···················· | ···················· | ···················· | NNNNNNNNNNNNNNNNNNNN | NNNNNNNNNNNNNNNNNNNN | 1 |
| L | 6896 hCoV-19/Belgium/ULG-10450/2020\|EPI_ISL_581625\|2020 | ···················· | ···················· | ···················· | ···················· | NNNNNNNNNNNNNNNNNNNN | NNNNNNNNNNNNNNNNNNNN | 1 |
| L | 6897 hCoV-19/Belgium/ULG-10451/2020\|EPI_ISL_581626\|2020 | ···················· | ···················· | ···················· | ···················· | NNNNNNNNNNNNNNNNNNNN | NNNNNNNNNNNNNNNNNNNN | 1 |
| L | 6899 hCoV-19/Belgium/ULG-10454/2020\|EPI_ISL_581628\|2020 | ···················· | ···················· | ···················· | ···················· | NNNNNNNNNNNNNNNNNNNN | NNNNNNNNNNNNNNNNNNNN | 1 |
| L | 6971 hCoV-19/Wales/PHWC-16B8FB/2020\|EPI_ISL_573916\|2020 | ···················· | ···················· | ···················· | ···················· | NNNNNNNNNNNNNNNNNNNN | NNNNNNNNNNNNNNNNNNNN | 1 |
| L | 6983 hCoV-19/Wales/PHWC-16B9F8/2020\|EPI_ISL_573928\|2020 | ···················· | ···················· | ···················· | ···················· | NNNNNNNNNNNNNNNNN··· | ···················· | 1 |
| L | 6995 hCoV-19/Wales/PHWC-16BAD7/2020\|EPI_ISL_573940\|2020 | ···················· | ···················· | ···················· | ···················· | NNNNNNNNNNNNNNNNNNNN | NNNNNNNNNNNNNNNNNNNN | 1 |
| L | 7002 hCoV-19/Wales/PHWC-16BB6B/2020\|EPI_ISL_573947\|2020 | ···················· | ···················· | ···················· | ···················· | NNNNNNNNNNNN········ | NNNNNNNNNNNNNNNNN··· | 1 |
| L | 7007 hCoV-19/Wales/PHWC-16BBC5/2020\|EPI_ISL_573952\|2020 | ···················· | ···················· | ···················· | ···················· | NNNNNNNNNNNNNNNNNNNN | ···················· | 1 |
| L | 7023 hCoV-19/Wales/PHWC-16BD56/2020\|EPI_ISL_573968\|2020 | ···················· | ···················· | ···················· | ···················· | NNNNNNNNNNNNNNNNNNNN | ···················· | 1 |
| L | 7045 hCoV-19/Wales/PHWC-16BF32/2020\|EPI_ISL_573990\|2020 | ···················· | ···················· | ···················· | ···················· | NNNNNNNNNNNNN······· | ···················· | 1 |
| L | 7047 hCoV-19/Wales/PHWC-16BF50/2020\|EPI_ISL_573992\|2020 | ···················· | ···················· | ···················· | ···················· | NNNN················ | ···················· | 1 |
| L | 7159 hCoV-19/USA/MI-UM-10035754891/2020\|EPI_ISL_583300\| | ···················· | ·A·················· | ···················· | ···················· | NNNNNNNNNNNNNNNNNNNN | ···················· | 1 |
| L | 7165 hCoV-19/USA/MI-UM-10035750305/2020\|EPI_ISL_583306\| | ···················· | ·············A······ | ···················· | ···················· | NNNNNNNNNNNNNNNNNNNN | ···················· | 1 |
| L | 7252 hCoV-19/Wales/PHWC-16D932/2020\|EPI_ISL_577467\|2020 | ···················· | ···················· | ···················· | ···················· | NNNNNNNNNNNNNNNNNNNN | NNNNNNNNNNNNNNNNNNNN | 1 |
| L | 7254 hCoV-19/Wales/PHWC-16D950/2020\|EPI_ISL_577469\|2020 | ···················· | ···················· | ···················· | ···················· | NNNNNNNNNNNNNNNNNNNN | NNNNNNNNNNNNNNNNNNNN | 1 |
| L | 7269 hCoV-19/Wales/PHWC-16DB3C/2020\|EPI_ISL_577494\|2020 | ···················· | ···················· | ···················· | ···················· | NNNN················ | ···················· | 1 |
| L | 7270 hCoV-19/Wales/PHWC-16DB4B/2020\|EPI_ISL_577495\|2020 | ···················· | ·A·······A·········· | ···················· | ···················· | NNNNNNNNNNNNNNNNNNNN | ···················· | 1 |
| L | 7274 hCoV-19/Wales/PHWC-16DB87/2020\|EPI_ISL_577499\|2020 | ···················· | ···················· | ···················· | ···················· | ·NNNNNNNNNNNNN······ | NNNN················ | 1 |
| L | 7276 hCoV-19/Wales/PHWC-16DBA5/2020\|EPI_ISL_577501\|2020 | ···················· | ···················· | ···················· | ···················· | NNNNNNNNNNNNNNNNNNNN | NNNNNNNNNNNNNNNNNNNN | 1 |
| L | 7278 hCoV-19/Wales/PHWC-16DBC3/2020\|EPI_ISL_577503\|2020 | ···················· | ···················· | ···················· | ···················· | NNN····NNNN········· | NNNNNNNNNNNNNNN····· | 1 |
| L | 7279 hCoV-19/Wales/PHWC-16DBD2/2020\|EPI_ISL_577504\|2020 | ···················· | ···················· | ···················· | ···················· | NNNNNNNNNNNNNNNNNNNN | ···················· | 1 |
| L | 7280 hCoV-19/Wales/PHWC-16DBE1/2020\|EPI_ISL_577505\|2020 | ···················· | ···················· | ···················· | ···················· | NNNNNNNNNNNNNNNNNNNN | ···················· | 1 |
| L | 7283 hCoV-19/Wales/PHWC-16DC1B/2020\|EPI_ISL_577508\|2020 | ···················· | ···················· | ···················· | ···················· | NNNNNNNNNNNNNNNNNNNN | ···················· | 1 |
| L | 7284 hCoV-19/Wales/PHWC-16DC2A/2020\|EPI_ISL_577509\|2020 | ···················· | ···················· | ···················· | ···················· | NNNNNNNNNNNNNNNNNNNN | ···················· | 1 |
| L | 7286 hCoV-19/Wales/PHWC-16DC57/2020\|EPI_ISL_577512\|2020 | ···················· | ···················· | ···················· | ···················· | NNNNNNNNNNNNNNNNNNNN | NNNNNNNNNNNNNNNNNNNN | 1 |
| L | 7287 hCoV-19/Wales/PHWC-16DC66/2020\|EPI_ISL_577513\|2020 | ···················· | ···················· | ···················· | ···················· | ········NNNNNNNNNNNN | ···················· | 1 |
| L | 7288 hCoV-19/Wales/PHWC-16DC84/2020\|EPI_ISL_577515\|2020 | ···················· | ···················· | ···················· | ···················· | ···········NNNNNNNNN | ···················· | 1 |
| L | 7290 hCoV-19/Wales/PHWC-16DCB1/2020\|EPI_ISL_577518\|2020 | ···················· | ···················· | ···················· | ···················· | NNNNNNNNNNNNNNNNNNNN | ···················· | 1 |
| L | 7291 hCoV-19/Wales/PHWC-16DCC0/2020\|EPI_ISL_577519\|2020 | ···················· | ···················· | ···················· | ···················· | NNNNNNNNNNNNNNNNNNNN | NNNNNNNNNNNNNNNNNNNN | 1 |
| L | 7292 hCoV-19/Wales/PHWC-16DCDF/2020\|EPI_ISL_577520\|2020 | ···················· | ···················· | ···················· | ···················· | NNNNNNNNNNNNNNNNNNNN | ···················· | 1 |
| L | 7293 hCoV-19/Wales/PHWC-16DCEE/2020\|EPI_ISL_577521\|2020 | ···················· | ···················· | ···················· | ···················· | NNNNNNNNNNNNNNNNNNNN | ···················· | 1 |
| L | 7294 hCoV-19/Wales/PHWC-16DCFD/2020\|EPI_ISL_577522\|2020 | ···················· | ···················· | ···················· | ···················· | NNNNNNNNNNNNNNNNNNNN | ···················· | 1 |
| L | 7295 hCoV-19/Wales/PHWC-16DD09/2020\|EPI_ISL_577523\|2020 | ···················· | ···················· | ···················· | ···················· | NNNNNNNNNNNNNNNNNNNN | ··NNNNNNNNNNNNNNNNNN | 1 |
| L | 7298 hCoV-19/Wales/PHWC-16DD36/2020\|EPI_ISL_577526\|2020 | ···················· | ···················· | ···················· | ···················· | NNNNNNNNNNNNNNNNNNNN | ···················· | 1 |
| L | 7299 hCoV-19/Wales/PHWC-16DD45/2020\|EPI_ISL_577527\|2020 | ···················· | ···················· | ···················· | ···················· | NNNNNNNNNNNNNNNNNNNN | ···················· | 1 |
| L | 7302 hCoV-19/Wales/PHWC-16DD72/2020\|EPI_ISL_577530\|2020 | ···················· | ···················· | ···················· | ···················· | NNNNNNNNNNNNNNNNNNNN | ···················· | 1 |
| L | 7306 hCoV-19/Wales/PHWC-16DDBE/2020\|EPI_ISL_577534\|2020 | ···················· | ···················· | ···················· | ···················· | NNNNNNNNNNNNNNNNNNNN | ···················· | 1 |
| L | 7308 hCoV-19/Wales/PHWC-16DE15/2020\|EPI_ISL_577540\|2020 | ···················· | ···················· | ···················· | ···················· | NNNNNNNNNNNNNNNNNNNN | ···················· | 1 |
| L | 7309 hCoV-19/Wales/PHWC-16DE24/2020\|EPI_ISL_577541\|2020 | ···················· | ···················· | ···················· | ···················· | NNNNNNNNNNNNNNNNNNNN | ···················· | 1 |
| L | 7310 hCoV-19/Wales/PHWC-16DE33/2020\|EPI_ISL_577542\|2020 | ···················· | ···················· | ···················· | ···················· | NNNNNNNNNNNNNNNNNNNN | ···················· | 1 |
| L | 7417 hCoV-19/Spain/PV-IBV-98007708/2020\|EPI_ISL_582085\| | ···················· | ···················· | ···················· | ···················· | ·NNNNNN············· | ··············N····· | 1 |
| L | 7424 hCoV-19/USA/MI-UM-10035921319/2020\|EPI_ISL_583399\| | ···················· | ···················· | ···················· | ···················· | NNNNNNNNNNNNNNNNNNNN | ···················· | 1 |
| L | 7570 hCoV-19/Wales/PHWC-16DDEB/2020\|EPI_ISL_577537\|2020 | ···················· | ···················· | ···················· | ···················· | NNNNNNNNNNNNNNNNNNNN | ···················· | 1 |
| L | 7575 hCoV-19/Switzerland/BS-UHB-65255924/2020\|EPI_ISL_5 | ···················· | ·A·················· | ···················· | ···················· | NNNNNNNNNNNNNNNNNNNN | ···················· | 1 |
| L | 7576 hCoV-19/Switzerland/BS-UHB-65256329/2020\|EPI_ISL_5 | ···················· | ···················· | ···················· | ···················· | NNNNNNNNNNNNNNNNNNNN | ···················· | 1 |
| L | 7618 hCoV-19/USA/MI-UM-10035787821/2020\|EPI_ISL_583323\| | ···················· | ·A·················· | ···················· | ···················· | NNNNNNNNNNNNNNNNNNNN | ···················· | 1 |
| L | 7623 hCoV-19/USA/MI-UM-10035862042/2020\|EPI_ISL_583328\| | T··················· | ·A·················· | ···················· | ···················· | NNNNNNNNNNNNNNNNNNNN | NNNNNNNNNNNNNNNNNNNN | 1 |
| L | 7625 hCoV-19/USA/MI-UM-10035862473/2020\|EPI_ISL_583330\| | ···················· | ·A·················· | ···················· | ···················· | NNNNNNNNNNNNNNNNNNNN | ···················· | 1 |
| L | 7628 hCoV-19/USA/MI-UM-10035853858/2020\|EPI_ISL_583333\| | ···················· | ·A·················· | ···················· | ···················· | NNNNNNNNNNNNNNNNNNNN | NNNNNNNNNNNNNNNNNNNN | 1 |
| L | 7629 hCoV-19/USA/MI-UM-10035868153/2020\|EPI_ISL_583334\| | ···················· | ···················· | ···················· | ···················· | NNNNNNNNNNNNNNNNNNNN | ···················· | 1 |
| L | 7630 hCoV-19/USA/MI-UM-10035861816/2020\|EPI_ISL_583335\| | T··················· | ·A·················· | ···················· | ···················· | NNNNNNNNNNNNNNNNNNNN | NNNNNNNNNNNNNNNNNNNN | 1 |
| L | 7639 hCoV-19/USA/MI-UM-10035872711/2020\|EPI_ISL_583346\| | T··················· | ·A·················· | ···················· | ···················· | NNNNNNNNNNNNNNNNNNNN | NNNNNNNNNNNNNNNNNNNN | 1 |
| L | 7640 hCoV-19/USA/MI-UM-10035955997/2020\|EPI_ISL_583347\| | ···················· | ···················· | ···················· | ···················· | NNNNNNNNNNNNNNNNNNNN | NNNNNNNNNNNNNNNNNNNN | 1 |
| L | 7648 hCoV-19/USA/MI-UM-10035981002/2020\|EPI_ISL_583357\| | ···················· | ···················· | ···················· | ···················· | NNNNNNNNNNNNNNNNNNNN | NNNNNNNNNNNNNNNNNNNN | 1 |
| L | 7649 hCoV-19/USA/MI-UM-10035988472/2020\|EPI_ISL_583360\| | ···················· | ·A·················· | ···················· | ···················· | NNNNNNNNNNNNNNNNNNNN | NNNNNNNNNNNNNNNNNNNN | 1 |
| L | 7651 hCoV-19/USA/MI-UM-10036001658/2020\|EPI_ISL_583363\| | ···················· | ···················· | ···················· | ···················· | NNNNNNNNNNNNNNNNNNNN | ···················· | 1 |
| L | 7654 hCoV-19/USA/MI-UM-10035968160/2020\|EPI_ISL_583366\| | ···················· | ···················· | ···················· | ···················· | NNNNNNN············· | ···················· | 1 |
| L | 7657 hCoV-19/USA/MI-UM-10036001045/2020\|EPI_ISL_583371\| | ···················· | ···················· | ···················· | ···················· | NNNNNNNNNNNNNNNNNNNN | NNNNNNNNNNNNNNNNNNNN | 1 |
| L | 7658 hCoV-19/USA/MI-UM-10036002831/2020\|EPI_ISL_583372\| | ···················· | ·A·················· | ···················· | ···················· | NNNNNNNNNNNNNNNNNNNN | ···················· | 1 |
| L | 7659 hCoV-19/USA/MI-UM-10035994708/2020\|EPI_ISL_583373\| | ···················· | ···················· | ···················· | ···················· | NNNNNNNNNNNNNNNNNNNN | ···················· | 1 |
| L | 7660 hCoV-19/USA/MI-UM-10036026125/2020\|EPI_ISL_583375\| | ···················· | ······A······A······ | ···················· | ···················· | NNNNNNNNNNNNNNNNNNNN | NNNNNNNNNNNNNNNNNNNN | 1 |
| L | 7662 hCoV-19/USA/MI-UM-10036033548/2020\|EPI_ISL_583377\| | ···················· | ···················· | ···················· | ···················· | NNNNNNNNNNNNNNNNNNNN | ···················· | 1 |
| L | 7663 hCoV-19/USA/MI-UM-10036004716/2020\|EPI_ISL_583378\| | ···················· | ·A·················· | ···················· | ···················· | NNNNNNNNNNNNNNNNNNNN | NNNNNNNNNNNNNNNNNNNN | 1 |
| L | 7667 hCoV-19/USA/MI-UM-10036037083/2020\|EPI_ISL_583383\| | ···················· | ······A······A······ | ···················· | ···················· | NNNNNNNNNNNNNNNNNNNN | NNNNNNNNNNNNNNNNNNNN | 1 |
| L | 7669 hCoV-19/USA/MI-UM-10036005251/2020\|EPI_ISL_583385\| | ···················· | ···················· | ···················· | ···················· | NNNNNNNNNNNNNNNNNNNN | NNNNNNNNNNNNNNNNNNNN | 1 |
| L | 7672 hCoV-19/USA/MI-UM-10035942254/2020\|EPI_ISL_583388\| | ···················· | ···················· | ···················· | ···················· | NNNNNNNNNNNNNNNNNNNN | ···················· | 1 |
| L | 7673 hCoV-19/USA/MI-UM-10035945885/2020\|EPI_ISL_583389\| | ···················· | ·············A······ | ···················· | ···················· | NNNNNNNNNNNNNNNNNNNN | ···················· | 1 |
| L | 7716 hCoV-19/Wales/MILK-9C4A98/2020\|EPI_ISL_573870\|2020 | ···················· | ···················· | ···················· | ···················· | NNNNNNNNNNNNNNNNNNNN | NNNNNNNNNNNNNNNNNNNN | 1 |
| L | 7781 hCoV-19/Scotland/EDB7776/2020\|EPI_ISL_577347\|2020- | ···················· | ···················· | ···················· | ···················· | NNNNNNNNNNNNNNNNNNNN | NNNNNNNNNNNNNNNNNNNN | 1 |
| L | 7782 hCoV-19/Scotland/EDB7785/2020\|EPI_ISL_577352\|2020- | ···················· | ······C············· | ···················· | ···················· | NNNNNNNNNNNNNNNNNNNN | NNNNNNNNNNNNNNNNNNNN | 1 |
| L | 7783 hCoV-19/Scotland/EDB7791/2020\|EPI_ISL_577354\|2020- | ···················· | ···················· | ···················· | ···················· | NNNNNNNNNNNNNNNNNNNN | NNNNNNNNNNNNNNNNNNNN | 1 |
| L | 7784 hCoV-19/Scotland/EDB7795/2020\|EPI_ISL_577357\|2020- | ···················· | ···················· | ···················· | ···················· | NNNNNNNNNNNNNNNNNNNN | NNNNNNNNNNNNNNNNNNNN | 1 |
| L | 7785 hCoV-19/Scotland/EDB7797/2020\|EPI_ISL_577358\|2020- | ···················· | ···················· | ···················· | ···················· | NNNNNNNNNNNNNNNNNNNN | NNNNNNNNNNNNNNNNNNNN | 1 |
| L | 7787 hCoV-19/Scotland/QEUH-9AE5B1/2020\|EPI_ISL_577360\|2 | ···················· | ···················· | ···················· | ···················· | NNNNNNNNNNNNNNNNNNNN | NNNNNNNNNNNNNNNNNNNN | 1 |
| L | 7809 hCoV-19/USA/WI-WSLH-200505/2020\|EPI_ISL_578646\|202 | ···················· | ···················· | ···················· | ···················· | NNNNNNNNNNNNNNNNNNNN | NNNNNNNNNNNNNNNNNNNN | 1 |
| L | 7810 hCoV-19/USA/WI-WSLH-200506/2020\|EPI_ISL_578647\|202 | ···················· | ···················· | ···················· | ···················· | NNNNNNNNNNNNNNNNNNNN | NNNNNNNNNNNNNNNNNNNN | 1 |
| L | 7811 hCoV-19/USA/WI-WSLH-200507/2020\|EPI_ISL_578648\|202 | ···················· | ···················· | ···················· | ···················· | NNNNNNNNNNNNNNNNNNNN | NNNNNNNNNNNNNNNNNNNN | 1 |

| | | | | | | | | | |
|---|---|---|---|---|---|---|---|---|---|
| L | 7812 | hCoV-19/USA/WI-WSLH-200508/2020\|EPI_ISL_578649\|202 | ···················· | ···················· | ···················· | ···················· | NNNNNNNNNNNNNNNNNNNN | NNNNNNNNNNNNNNNNNNNN | 1 |
| L | 7813 | hCoV-19/USA/WI-WSLH-200509/2020\|EPI_ISL_578650\|202 | ···················· | ···················· | ···················· | ···················· | NNNNNNNNNNNNNNNNNNNN | NNNNNNNNNNNNNNNNNNNN | 1 |
| L | 7814 | hCoV-19/USA/WI-WSLH-200510/2020\|EPI_ISL_578651\|202 | ···················· | ···················· | ···················· | ···················· | NNNNNNNNNNNNNNNNNNNN | NNNNNNNNNNNNNNNNNNNN | 1 |
| L | 7815 | hCoV-19/USA/WI-WSLH-200511/2020\|EPI_ISL_578652\|202 | ···················· | ···················· | ···················· | ···················· | NNNNNNNNNNNNNNNNNNNN | NNNNNNNNNNNNNNNNNNNN | 1 |
| L | 7818 | hCoV-19/USA/WI-WSLH-200514/2020\|EPI_ISL_578655\|202 | ···················· | ···················· | ···················· | ···················· | NNNNNNNNNNNNNNNNNNNN | NNNNNNNNNNNNNNNNNNNN | 1 |
| L | 7819 | hCoV-19/USA/WI-WSLH-200516/2020\|EPI_ISL_578656\|202 | ···················· | ···················· | ···················· | ···················· | NNNNNNNNNNNNNNNNNNNN | NNNNNNNNNNNNNNNNNNNN | 1 |
| L | 7839 | hCoV-19/USA/WI-WSLH-200556/2020\|EPI_ISL_578676\|202 | ···················· | ···················· | ···················· | ···················· | NNNNNNNNNNNNNNNNNNNN | NNNNNNNNNNNNNNNNNNNN | 1 |
| L | 7840 | hCoV-19/USA/WI-WSLH-200559/2020\|EPI_ISL_578677\|202 | ···················· | ···················· | ···················· | ···················· | NNNNNNNNNNNNNNNNNNNN | NNNNNNNNNNNNNNNNNNNN | 1 |
| L | 7841 | hCoV-19/USA/WI-WSLH-200565/2020\|EPI_ISL_578678\|202 | ···················· | ···················· | ···················· | ···················· | NNNNNNNNNNNNNNNNNNNN | NNNNNNNNNNNNNNNNNNNN | 1 |
| L | 7842 | hCoV-19/USA/WI-WSLH-200566/2020\|EPI_ISL_578679\|202 | ···················· | ···················· | ···················· | ···················· | NNNNNNNNNNNNNNNNNNNN | NNNNNNNNNNNNNNNNNNNN | 1 |
| L | 7843 | hCoV-19/USA/WI-WSLH-200568/2020\|EPI_ISL_578680\|202 | ···················· | ···················· | ···················· | ···················· | NNNNNNNNNNNNNNNNNNNN | NNNNNNNNNNNNNNNNNNNN | 1 |
| L | 7845 | hCoV-19/USA/WI-WSLH-200570/2020\|EPI_ISL_578682\|202 | ···················· | ···················· | ···················· | ···················· | NNNNNNNNNNNNNNNNNNNN | NNNNNNNNNNNNNNNNNNNN | 1 |
| L | 7846 | hCoV-19/USA/WI-WSLH-200571/2020\|EPI_ISL_578683\|202 | ···················· | ···················· | ···················· | ···················· | NNNNNNNNNNNNNNNNNNNN | NNNNNNNNNNNNNNNNNNNN | 1 |
| L | 7847 | hCoV-19/USA/WI-WSLH-200572/2020\|EPI_ISL_578684\|202 | ···················· | ···················· | ···················· | ···················· | NNNNNNNNNNNNNNNNNNNN | NNNNNNNNNNNNNNNNNNNN | 1 |
| L | 7848 | hCoV-19/USA/WI-WSLH-200573/2020\|EPI_ISL_578685\|202 | ···················· | ···················· | ···················· | ···················· | NNNNNNNNNNNNNNNNNNNN | NNNNNNNNNNNNNNNNNNNN | 1 |
| L | 7849 | hCoV-19/USA/WI-WSLH-200575/2020\|EPI_ISL_578686\|202 | ···················· | ···················· | ···················· | ···················· | NNNNNNNNNNNNNNNNNNNN | NNNNNNNNNNNNNNNNNNNN | 1 |
| L | 7850 | hCoV-19/USA/WI-WSLH-200576/2020\|EPI_ISL_578687\|202 | ···················· | ···················· | ···················· | ···················· | NNNNNNNNNNNNNNNNNNNN | NNNNNNNNNNNNNNNNNNNN | 1 |
| L | 7851 | hCoV-19/USA/WI-WSLH-200577/2020\|EPI_ISL_578688\|202 | ···················· | ···················· | ···················· | ···················· | NNNNNNNNNNNNNNNNNNNN | NNNNNNNNNNNNNNNNNNNN | 1 |
| L | 7852 | hCoV-19/USA/WI-WSLH-200578/2020\|EPI_ISL_578689\|202 | ···················· | ···················· | ···················· | ···················· | NNNNNNNNNNNNNNNNNNNN | NNNNNNNNNNNNNNNNNNNN | 1 |
| L | 7853 | hCoV-19/USA/WI-WSLH-200579/2020\|EPI_ISL_578690\|202 | ···················· | ···················· | ···················· | ···················· | NNNNNNNNNNNNNNNNNNNN | NNNNNNNNNNNNNNNNNNNN | 1 |
| L | 7854 | hCoV-19/USA/WI-WSLH-200580/2020\|EPI_ISL_578691\|202 | ···················· | ···················· | ···················· | ···················· | NNNNNNNNNNNNNNNNNNNN | NNNNNNNNNNNNNNNNNNNN | 1 |
| L | 7856 | hCoV-19/USA/WI-WSLH-200582/2020\|EPI_ISL_578693\|202 | ···················· | ···················· | ···················· | ···················· | NNNNNNNNNNNNNNNNNNNN | NNNNNNNNNNNNNNNNNNNN | 1 |
| L | 7857 | hCoV-19/USA/WI-WSLH-200584/2020\|EPI_ISL_578694\|202 | ···················· | ···················· | ···················· | ···················· | NNNNNNNNNNNNNNNNNNNN | NNNNNNNNNNNNNNNNNNNN | 1 |
| L | 7858 | hCoV-19/USA/WI-WSLH-200585/2020\|EPI_ISL_578695\|202 | ···················· | ···················· | ···················· | ···················· | NNNNNNNNNNNNNNNNNNNN | NNNNNNNNNNNNNNNNNNNN | 1 |
| L | 7936 | hCoV-19/USA/MI-UM-10035681090/2020\|EPI_ISL_583287\| | ···················· | ···················· | ···················· | ···················· | NNNNNNNNNNNNNNNNNNNN | NNNNNNNNNNNNNNNNNNNN | 1 |
| L | 8074 | hCoV-19/Wales/PHWC-16D6B3/2020\|EPI_ISL_577431\|2020 | ···················· | ···················· | ···················· | ···················· | NNNNNNNNNNNNNNNNNNNN | NNNNNNNNNNNNNNNNNNNN | 1 |
| L | 8162 | hCoV-19/USA/MI-UM-10035702357/2020\|EPI_ISL_583249\| | ···················· | ···················· | ···················· | ···················· | NNNNNNNNNNNNNNNNNNNN | ···················· | 1 |
| L | 8163 | hCoV-19/USA/MI-UM-10035713127/2020\|EPI_ISL_583255\| | ···················· | ·············A······ | ···················· | ···················· | NNNNNNNNNNNNNNNNNNNN | NNNNNNNNNNNNNNNNNNNN | 1 |
| L | 8164 | hCoV-19/USA/MI-UM-10035685450/2020\|EPI_ISL_583257\| | ···················· | ·A·················· | ···················· | ···················· | NNNNNNNNNNNNNNNNNNNN | NNNNNNNNNNNNNNNNNNNN | 1 |
| L | 8165 | hCoV-19/USA/MI-UM-10035547701/2020\|EPI_ISL_583258\| | ···················· | ···················· | ···················· | ···················· | NNNNNNNNNNNNNNNNNNNN | NNNNNNNNNNNNNNNNNNNN | 1 |
| L | 8166 | hCoV-19/USA/MI-UM-10035683340/2020\|EPI_ISL_583259\| | ···················· | ···················· | ···················· | ···················· | NNNNNNNNNNNNNNNNNNNN | NNNNNNNNNNNNNNNNNNNN | 1 |
| L | 8168 | hCoV-19/USA/MI-UM-10035638988/2020\|EPI_ISL_583261\| | ···················· | ···················· | ···················· | ···················· | NNNNNNNNNNNNNNNNNNNN | ···················· | 1 |
| L | 8169 | hCoV-19/USA/MI-UM-10035692188/2020\|EPI_ISL_583262\| | ···················· | ·A·················· | ···················· | ···················· | NNNNNNNNNNNNNNNNNNNN | NNNNNNNNNNNNNNNNNNNN | 1 |
| L | 8173 | hCoV-19/USA/MI-UM-10035665912/2020\|EPI_ISL_583267\| | ···················· | ···················· | ···················· | ···················· | NNNNNNNNNNNNNNNNNNNN | ···················· | 1 |
| L | 8174 | hCoV-19/USA/MI-UM-10035696298/2020\|EPI_ISL_583268\| | ···················· | ···················· | ···················· | ···················· | NNNNNNNNNNNNNNNNNNNN | ···················· | 1 |
| L | 8178 | hCoV-19/USA/MI-UM-10035685395/2020\|EPI_ISL_583272\| | ···················· | ···················· | ···················· | ···················· | ··NNNNNNNNNNNNNNNNNN | ···················· | 1 |
| L | 8179 | hCoV-19/USA/MI-UM-10035701578/2020\|EPI_ISL_583273\| | ···················· | ·A·················· | ···················· | ···················· | NNNNNNNNNNNNNNNNNNNN | NNNNNNNNNNNNNNNNNNNN | 1 |
| L | 8185 | hCoV-19/USA/MI-UM-10035772246/2020\|EPI_ISL_583290\| | ···················· | ···················· | ···················· | ···················· | NNNNNNNNNNNNNNNNNNNN | ···················· | 1 |
| L | 8186 | hCoV-19/USA/MI-UM-10035755495/2020\|EPI_ISL_583292\| | ···················· | ···················· | ···················· | ···················· | NNNNNNNNNNNNNNNNNNNN | ···················· | 1 |
| L | 8192 | hCoV-19/USA/MI-UM-10035734995/2020\|EPI_ISL_583298\| | ···················· | ···················· | ···················· | ···················· | NNNNNNNNNNNNNNNNNNNN | ···················· | 1 |
| L | 8255 | hCoV-19/England/QEUH-9AE733/2020\|EPI_ISL_577235\|20 | ···················· | ···················· | ···················· | ···················· | NNNNNNNNNNNNNNNNNNNN | NNNNNNNNNNNNNNNNNNNN | 1 |
| L | 8260 | hCoV-19/England/QEUH-9AE812/2020\|EPI_ISL_577244\|20 | ···················· | ···················· | ···················· | ···················· | NNNNNNNNNNNNNNNNNNNN | NNNNNNNNNNNNNNNNNNNN | 1 |
| L | 8265 | hCoV-19/England/QEUH-9AEA3A/2020\|EPI_ISL_577267\|20 | ···················· | ···················· | ···················· | ···················· | NNNNNNNNNNNNNNNNNNNN | ···················· | 1 |
| L | 8267 | hCoV-19/England/QEUH-9AEA85/2020\|EPI_ISL_577271\|20 | ···················· | ···················· | ···················· | ···················· | NNNNNNNNNNNNNNNNNNNN | NNNNNNNNNNNNNNNNNNNN | 1 |
| L | 8281 | hCoV-19/England/QEUH-9D15A6/2020\|EPI_ISL_577290\|20 | ···················· | NNNNNNNNNNNNNNNNNNNN | ···················· | ···················· | ···················· | ···················· | 1 |
| L | 8300 | hCoV-19/USA/WI-WSLH-200367/2020\|EPI_ISL_578536\|202 | ···················· | ···················· | ···················· | ···················· | NNNNNNNNNNNNNNNNNNNN | NNNNNNNNNNNNNNNNNNNN | 1 |
| L | 8309 | hCoV-19/USA/WI-WSLH-200376/2020\|EPI_ISL_578545\|202 | ···················· | ···················· | ···················· | ···················· | NNNNNNNNNNNNNNNNNNNN | NNNNNNNNNNNNNNNNNNNN | 1 |
| L | 8311 | hCoV-19/USA/WI-WSLH-200379/2020\|EPI_ISL_578547\|202 | ···················· | ···················· | ···················· | ···················· | NNNNNNNNNNNNNNNNNNNN | NNNNNNNNNNNNNNNNNNNN | 1 |
| L | 8312 | hCoV-19/USA/WI-WSLH-200380/2020\|EPI_ISL_578548\|202 | ···················· | ···················· | ···················· | ···················· | NNNNNNNNNNNNNNNNNNNN | NNNNNNNNNNNNNNNNNNNN | 1 |
| L | 8313 | hCoV-19/USA/WI-WSLH-200381/2020\|EPI_ISL_578549\|202 | ···················· | ···················· | ···················· | ···················· | NNNNNNNNNNNNNNNNNNNN | NNNNNNNNNNNNNNNNNNNN | 1 |
| L | 8314 | hCoV-19/USA/WI-WSLH-200383/2020\|EPI_ISL_578550\|202 | ···················· | ···················· | ···················· | ···················· | NNNNNNNNNNNNNNNNNNNN | NNNNNNNNNNNNNNNNNNNN | 1 |
| L | 8315 | hCoV-19/USA/WI-WSLH-200384/2020\|EPI_ISL_578551\|202 | ···················· | ···················· | ···················· | ···················· | NNNNNNNNNNNNNNNNNNNN | NNNNNNNNNNNNNNNNNNNN | 1 |
| L | 8316 | hCoV-19/USA/WI-WSLH-200386/2020\|EPI_ISL_578552\|202 | ···················· | ···················· | ···················· | ···················· | NNNNNNNNNNNNNNNNNNNN | NNNNNNNNNNNNNNNNNNNN | 1 |
| L | 8317 | hCoV-19/USA/WI-WSLH-200387/2020\|EPI_ISL_578553\|202 | ···················· | ···················· | ···················· | ···················· | NNNNNNNNNNNNNNNNNNNN | NNNNNNNNNNNNNNNNNNNN | 1 |
| L | 8318 | hCoV-19/USA/WI-WSLH-200388/2020\|EPI_ISL_578554\|202 | ···················· | ···················· | ···················· | ···················· | NNNNNNNNNNNNNNNNNNNN | NNNNNNNNNNNNNNNNNNNN | 1 |
| L | 8319 | hCoV-19/USA/WI-WSLH-200389/2020\|EPI_ISL_578555\|202 | ···················· | ···················· | ···················· | ···················· | NNNNNNNNNNNNNNNNNNNN | NNNNNNNNNNNNNNNNNNNN | 1 |
| L | 8320 | hCoV-19/USA/WI-WSLH-200390/2020\|EPI_ISL_578556\|202 | ···················· | ···················· | ···················· | ···················· | NNNNNNNNNNNNNNNNNNNN | NNNNNNNNNNNNNNNNNNNN | 1 |
| L | 8321 | hCoV-19/USA/WI-WSLH-200391/2020\|EPI_ISL_578557\|202 | ···················· | ···················· | ···················· | ···················· | NNNNNNNNNNNNNNNNNNNN | NNNNNNNNNNNNNNNNNNNN | 1 |
| L | 8322 | hCoV-19/USA/WI-WSLH-200392/2020\|EPI_ISL_578558\|202 | ···················· | ···················· | ···················· | ···················· | NNNNNNNNNNNNNNNNNNNN | NNNNNNNNNNNNNNNNNNNN | 1 |
| L | 8323 | hCoV-19/USA/WI-WSLH-200393/2020\|EPI_ISL_578559\|202 | ···················· | ···················· | ···················· | ···················· | NNNNNNNNNNNNNNNNNNNN | NNNNNNNNNNNNNNNNNNNN | 1 |
| L | 8324 | hCoV-19/USA/WI-WSLH-200394/2020\|EPI_ISL_578560\|202 | ···················· | ···················· | ···················· | ···················· | NNNNNNNNNNNNNNNNNNNN | NNNNNNNNNNNNNNNNNNNN | 1 |
| L | 8325 | hCoV-19/USA/WI-WSLH-200395/2020\|EPI_ISL_578561\|202 | ···················· | ···················· | ···················· | ···················· | NNNNNNNNNNNNNNNNNNNN | NNNNNNNNNNNNNNNNNNNN | 1 |
| L | 8326 | hCoV-19/USA/WI-WSLH-200396/2020\|EPI_ISL_578562\|202 | ···················· | ···················· | ···················· | ···················· | NNNNNNNNNNNNNNNNNNNN | NNNNNNNNNNNNNNNNNNNN | 1 |
| L | 8327 | hCoV-19/USA/WI-WSLH-200397/2020\|EPI_ISL_578563\|202 | ···················· | ···················· | ···················· | ···················· | NNNNNNNNNNNNNNNNNNNN | NNNNNNNNNNNNNNNNNNNN | 1 |
| L | 8328 | hCoV-19/USA/WI-WSLH-200398/2020\|EPI_ISL_578564\|202 | ···················· | ···················· | ···················· | ···················· | NNNNNNNNNNNNNNNNNNNN | NNNNNNNNNNNNNNNNNNNN | 1 |
| L | 8329 | hCoV-19/USA/WI-WSLH-200399/2020\|EPI_ISL_578565\|202 | ···················· | ···················· | ···················· | ···················· | NNNNNNNNNNNNNNNNNNNN | NNNNNNNNNNNNNNNNNNNN | 1 |
| L | 8330 | hCoV-19/USA/WI-WSLH-200400/2020\|EPI_ISL_578566\|202 | ···················· | ···················· | ···················· | ···················· | NNNNNNNNNNNNNNNNNNNN | NNNNNNNNNNNNNNNNNNNN | 1 |
| L | 8331 | hCoV-19/USA/WI-WSLH-200401/2020\|EPI_ISL_578567\|202 | ···················· | ···················· | ···················· | ···················· | NNNNNNNNNNNNNNNNNNNN | NNNNNNNNNNNNNNNNNNNN | 1 |
| L | 8332 | hCoV-19/USA/WI-WSLH-200402/2020\|EPI_ISL_578568\|202 | ···················· | ···················· | ···················· | ···················· | NNNNNNNNNNNNNNNNNNNN | NNNNNNNNNNNNNNNNNNNN | 1 |
| L | 8333 | hCoV-19/USA/WI-WSLH-200403/2020\|EPI_ISL_578569\|202 | ···················· | ···················· | ···················· | ···················· | NNNNNNNNNNNNNNNNNNNN | NNNNNNNNNNNNNNNNNNNN | 1 |
| L | 8334 | hCoV-19/USA/WI-WSLH-200405/2020\|EPI_ISL_578570\|202 | ···················· | ···················· | ···················· | ···················· | NNNNNNNNNNNNNNNNNNNN | NNNNNNNNNNNNNNNNNNNN | 1 |
| L | 8335 | hCoV-19/USA/WI-WSLH-200407/2020\|EPI_ISL_578571\|202 | ···················· | ···················· | ···················· | ···················· | NNNNNNNNNNNNNNNNNNNN | NNNNNNNNNNNNNNNNNNNN | 1 |
| L | 8336 | hCoV-19/USA/WI-WSLH-200408/2020\|EPI_ISL_578572\|202 | ···················· | ···················· | ···················· | ···················· | NNNNNNNNNNNNNNNNNNNN | NNNNNNNNNNNNNNNNNNNN | 1 |
| L | 8337 | hCoV-19/USA/WI-WSLH-200409/2020\|EPI_ISL_578573\|202 | ···················· | ···················· | ···················· | ···················· | NNNNNNNNNNNNNNNNNNNN | NNNNNNNNNNNNNNNNNNNN | 1 |
| L | 8338 | hCoV-19/USA/WI-WSLH-200411/2020\|EPI_ISL_578574\|202 | ···················· | ···················· | ···················· | ···················· | NNNNNNNNNNNNNNNNNNNN | NNNNNNNNNNNNNNNNNNNN | 1 |
| L | 8339 | hCoV-19/USA/WI-WSLH-200412/2020\|EPI_ISL_578575\|202 | ···················· | ···················· | ···················· | ···················· | NNNNNNNNNNNNNNNNNNNN | NNNNNNNNNNNNNNNNNNNN | 1 |
| L | 8340 | hCoV-19/USA/WI-WSLH-200414/2020\|EPI_ISL_578576\|202 | ·T·················· | ···················· | ···················· | ···················· | NNNNNNNNNNNNNNNNNNNN | NNNNNNNNNNNNNNNNNNNN | 1 |
| L | 8341 | hCoV-19/USA/WI-WSLH-200415/2020\|EPI_ISL_578577\|202 | ···················· | ···················· | ···················· | ···················· | NNNNNNNNNNNNNNNNNNNN | NNNNNNNNNNNNNNNNNNNN | 1 |
| L | 8342 | hCoV-19/USA/WI-WSLH-200416/2020\|EPI_ISL_578578\|202 | ···················· | ···················· | ···················· | ···················· | NNNNNNNNNNNNNNNNNNNN | NNNNNNNNNNNNNNNNNNNN | 1 |
| L | 8343 | hCoV-19/USA/WI-WSLH-200417/2020\|EPI_ISL_578579\|202 | ···················· | ···················· | ···················· | ···················· | NNNNNNNNNNNNNNNNNNNN | NNNNNNNNNNNNNNNNNNNN | 1 |

| | | | | | | | | |
|---|---|---|---|---|---|---|---|---|
| L | 8344 hCoV-19/USA/WI-WSLH-200418/2020\|EPI_ISL_578580\|202 | ···················· | ···················· | ···················· | ···················· | NNNNNNNNNNNNNNNNNNNN | NNNNNNNNNNNNNNNNNNNN | 1 |
| L | 8345 hCoV-19/USA/WI-WSLH-200419/2020\|EPI_ISL_578581\|202 | ···················· | ···················· | ···················· | ···················· | NNNNNNNNNNNNNNNNNNNN | NNNNNNNNNNNNNNNNNNNN | 1 |
| L | 8346 hCoV-19/USA/WI-WSLH-200420/2020\|EPI_ISL_578582\|202 | ···················· | ···················· | ···················· | ···················· | NNNNNNNNNNNNNNNNNNNN | NNNNNNNNNNNNNNNNNNNN | 1 |
| L | 8347 hCoV-19/USA/WI-WSLH-200421/2020\|EPI_ISL_578583\|202 | ···················· | ···················· | ···················· | ···················· | NNNNNNNNNNNNNNNNNNNN | NNNNNNNNNNNNNNNNNNNN | 1 |
| L | 8348 hCoV-19/USA/WI-WSLH-200423/2020\|EPI_ISL_578584\|202 | ···················· | ···················· | ···················· | ···················· | NNNNNNNNNNNNNNNNNNNN | NNNNNNNNNNNNNNNNNNNN | 1 |
| L | 8349 hCoV-19/USA/WI-WSLH-200426/2020\|EPI_ISL_578586\|202 | ···················· | ···················· | ···················· | ···················· | NNNNNNNNNNNNNNNNNNNN | NNNNNNNNNNNNNNNNNNNN | 1 |
| L | 8350 hCoV-19/USA/WI-WSLH-200428/2020\|EPI_ISL_578587\|202 | ···················· | ···················· | ···················· | ···················· | NNNNNNNNNNNNNNNNNNNN | NNNNNNNNNNNNNNNNNNNN | 1 |
| L | 8351 hCoV-19/USA/WI-WSLH-200434/2020\|EPI_ISL_578588\|202 | ···················· | ···················· | ···················· | ···················· | NNNNNNNNNNNNNNNNNNNN | NNNNNNNNNNNNNNNNNNNN | 1 |
| L | 8352 hCoV-19/USA/WI-WSLH-200440/2020\|EPI_ISL_578589\|202 | ···················· | ···················· | ···················· | ···················· | NNNNNNNNNNNNNNNNNNNN | NNNNNNNNNNNNNNNNNNNN | 1 |
| L | 8446 hCoV-19/USA/MI-UM-10035695135/2020\|EPI_ISL_583252\| | ···················· | ············A······· | ···················· | ···················· | NNNNNNNNNNNNNNNNNNNN | ···················· | 1 |
| L | 8447 hCoV-19/USA/MI-UM-10035685254/2020\|EPI_ISL_583253\| | ···················· | ···················· | ···················· | ···················· | NNNNNNNNNNNNNNNNNNNN | ···················· | 1 |
| L | 8480 hCoV-19/USA/WI-WSLH-200442/2020\|EPI_ISL_578590\|202 | ···················· | ···················· | ···················· | ···················· | NNNNNNNNNNNNNNNNNNNN | NNNNNNNNNNNNNNNNNNNN | 1 |
| L | 8482 hCoV-19/USA/WI-WSLH-200446/2020\|EPI_ISL_578592\|202 | ···················· | ···················· | ···················· | ···················· | NNNNNNNNNNNNNNNNNNNN | NNNNNNNNNNNNNNNNNNNN | 1 |
| L | 8484 hCoV-19/USA/WI-WSLH-200449/2020\|EPI_ISL_578594\|202 | ···················· | ···················· | ···················· | ···················· | NNNNNNNNNNNNNNNNNNNN | NNNNNNNNNNNNNNNNNNNN | 1 |
| L | 8486 hCoV-19/USA/WI-WSLH-200451/2020\|EPI_ISL_578596\|202 | ···················· | ···················· | ···················· | ···················· | NNNNNNNNNNNNNNNNNNNN | NNNNNNNNNNNNNNNNNNNN | 1 |
| L | 8499 hCoV-19/USA/WI-WSLH-200471/2020\|EPI_ISL_578612\|202 | ···················· | ···················· | ···················· | ···················· | NNNNNNNNNNNNNNNNNNNN | NNNNNNNNNNNNNNNNNNNN | 1 |
| L | 8504 hCoV-19/USA/WI-WSLH-200476/2020\|EPI_ISL_578617\|202 | ···················· | ···················· | ···················· | ···················· | NNNNNNNNNNNNNNNNNNNN | NNNNNNNNNNNNNNNNNNNN | 1 |
| L | 8505 hCoV-19/USA/WI-WSLH-200477/2020\|EPI_ISL_578618\|202 | ···················· | ···················· | ···················· | ···················· | NNNNNNNNNNNNNNNNNNNN | NNNNNNNNNNNNNNNNNNNN | 1 |
| L | 8507 hCoV-19/USA/WI-WSLH-200479/2020\|EPI_ISL_578620\|202 | ···················· | ···················· | ···················· | ···················· | NNNNNNNNNNNNNNNNNNNN | NNNNNNNNNNNNNNNNNNNN | 1 |
| L | 8508 hCoV-19/USA/WI-WSLH-200480/2020\|EPI_ISL_578621\|202 | ···················· | ···················· | ···················· | ···················· | NNNNNNNNNNNNNNNNNNNN | NNNNNNNNNNNNNNNNNNNN | 1 |
| L | 8509 hCoV-19/USA/WI-WSLH-200481/2020\|EPI_ISL_578622\|202 | ···················· | ···················· | ···················· | ···················· | NNNNNNNNNNNNNNNNNNNN | ···················· | 1 |
| L | 8510 hCoV-19/USA/WI-WSLH-200482/2020\|EPI_ISL_578623\|202 | ···················· | ···················· | ···················· | ···················· | NNNNNNNNNNNNNNNNNNNN | NNNNNNNNNNNNNNNNNNNN | 1 |
| L | 8511 hCoV-19/USA/WI-WSLH-200483/2020\|EPI_ISL_578624\|202 | ···················· | ···················· | ···················· | ···················· | NNNNNNNNNNNNNNNNNNNN | NNNNNNNNNNNNNNNNNNNN | 1 |
| L | 8512 hCoV-19/USA/WI-WSLH-200484/2020\|EPI_ISL_578625\|202 | ···················· | ···················· | ···················· | ···················· | NNNNNNNNNNNNNNNNNNNN | NNNNNNNNNNNNNNNNNNNN | 1 |
| L | 8515 hCoV-19/USA/WI-WSLH-200487/2020\|EPI_ISL_578628\|202 | ···················· | ···················· | ···················· | ···················· | NNNNNNNNNNNNNNNNNNNN | NNNNNNNNNNNNNNNNNNNN | 1 |
| L | 8516 hCoV-19/USA/WI-WSLH-200488/2020\|EPI_ISL_578629\|202 | ···················· | ···················· | ···················· | ···················· | NNNNNNNNNNNNNNNNNNNN | NNNNNNNNNNNNNNNNNNNN | 1 |
| L | 8518 hCoV-19/USA/WI-WSLH-200490/2020\|EPI_ISL_578631\|202 | ···················· | ···················· | ···················· | ···················· | NNNNNNNNNNNNNNNNNNNN | NNNNNNNNNNNNNNNNNNNN | 1 |
| L | 8519 hCoV-19/USA/WI-WSLH-200491/2020\|EPI_ISL_578632\|202 | ···················· | ···················· | ···················· | ···················· | NNNNNNNNNNNNNNNNNNNN | NNNNNNNNNNNNNNNNNNNN | 1 |
| L | 8520 hCoV-19/USA/WI-WSLH-200492/2020\|EPI_ISL_578633\|202 | ···················· | ···················· | ···················· | ···················· | NNNNNNNNNNNNNNNNNNNN | NNNNNNNNNNNNNNNNNNNN | 1 |
| L | 8522 hCoV-19/USA/WI-WSLH-200494/2020\|EPI_ISL_578635\|202 | ···················· | ···················· | ···················· | ···················· | NNNNNNNNNNNNNNNNNNNN | NNNNNNNNNNNNNNNNNNNN | 1 |
| L | 8523 hCoV-19/USA/WI-WSLH-200495/2020\|EPI_ISL_578636\|202 | ···················· | ···················· | ···················· | ···················· | NNNNNNNNNNNNNNNNNNNN | NNNNNNNNNNNNNNNNNNNN | 1 |
| L | 8524 hCoV-19/USA/WI-WSLH-200496/2020\|EPI_ISL_578637\|202 | ···················· | ···················· | ···················· | ···················· | NNNNNNNNNNNNNNNNNNNN | NNNNNNNNNNNNNNNNNNNN | 1 |
| L | 8525 hCoV-19/USA/WI-WSLH-200497/2020\|EPI_ISL_578638\|202 | ···················· | ···················· | ···················· | ···················· | NNNNNNNNNNNNNNNNNNNN | NNNNNNNNNNNNNNNNNNNN | 1 |
| L | 8526 hCoV-19/USA/WI-WSLH-200498/2020\|EPI_ISL_578639\|202 | ···················· | ···················· | ···················· | ···················· | NNNNNNNNNNNNNNNNNNNN | NNNNNNNNNNNNNNNNNNNN | 1 |
| L | 8529 hCoV-19/USA/WI-WSLH-200501/2020\|EPI_ISL_578642\|202 | ···················· | ···················· | ···················· | ···················· | NNNNNNNNNNNNNNNNNNNN | NNNNNNNNNNNNNNNNNNNN | 1 |
| L | 8530 hCoV-19/USA/WI-WSLH-200502/2020\|EPI_ISL_578643\|202 | ···················· | ···················· | ···················· | ···················· | NNNNNNNNNNNNNNNNNNNN | NNNNNNNNNNNNNNNNNNNN | 1 |
| L | 8531 hCoV-19/USA/WI-WSLH-200503/2020\|EPI_ISL_578644\|202 | ···················· | ···················· | ···················· | ···················· | NNNNNNNNNNNNNNNNNNNN | NNNNNNNNNNNNNNNNNNNN | 1 |
| L | 8532 hCoV-19/USA/WI-WSLH-200504/2020\|EPI_ISL_578645\|202 | ···················· | ···················· | ···················· | ···················· | NNNNNNNNNNNNNNNNNNNN | NNNNNNNNNNNNNNNNNNNN | 1 |
| L | 8666 hCoV-19/USA/CA-CZB-10932/2020\|EPI_ISL_583161\|2020- | ···················· | ·A·················· | ···················· | ···················· | NNNNNNNNNNN········· | NN·······NNNNNN····· | 1 |
| L | 8669 hCoV-19/USA/CA-CZB-10935/2020\|EPI_ISL_583164\|2020- | ···················· | ·A·················· | ···················· | ···················· | NNNNNNNNNNNNNNNNNNNN | NNNNNNNNNNNNNNNNNNNN | 1 |
| L | 8807 hCoV-19/Scotland/CVR4125/2020\|EPI_ISL_573767\|2020- | ···················· | ···················· | ···················· | ···················· | NNNNNNNNNNNNNNNNNNNN | NNNNNNNNNNNNNNNNNNNN | 1 |
| L | 8811 hCoV-19/Scotland/CVR4321/2020\|EPI_ISL_573772\|2020- | ···················· | ···················· | ···················· | ···················· | NNNNNNNNNNNNNNNNNNNN | NNNNNNNNNNNNNNNNNNNN | 1 |
| L | 8817 hCoV-19/Scotland/CVR4337/2020\|EPI_ISL_573778\|2020- | ···················· | ···················· | ···················· | ···················· | ···N·NNN············ | NNNN···NN····NN····· | 1 |
| L | 8834 hCoV-19/Scotland/EDB7819/2020\|EPI_ISL_573795\|2020- | ···················· | ···················· | ···················· | ···················· | NNNNNNNNNNNNNNNNNNNN | NNNNNNNNNNNNNNNNNNNN | 1 |
| L | 8838 hCoV-19/Scotland/MILK-9C72BB/2020\|EPI_ISL_573799\|2 | ···················· | ···················· | ···················· | ···················· | NNNNNNNNNNNNNNNNNNNN | NNNNNNNNNNNNNNNNNNNN | 1 |
| L | 8899 hCoV-19/USA/WI-WSLH-200308/2020\|EPI_ISL_578490\|202 | ···················· | ···················· | ···················· | ···················· | NNNNNNNNNNNNNNNNNNNN | NNNNNNNNNNNNNNNNNNNN | 1 |
| L | 8900 hCoV-19/USA/WI-WSLH-200310/2020\|EPI_ISL_578491\|202 | ···················· | ···················· | ···················· | ···················· | NNNNNNNNNNNNNNNNNNNN | NNNNNNNNNNNNNNNNNNNN | 1 |
| L | 8901 hCoV-19/USA/WI-WSLH-200311/2020\|EPI_ISL_578492\|202 | ···················· | ···················· | ···················· | ···················· | NNNNNNNNNNNNNNNNNNNN | NNNNNNNNNNNNNNNNNNNN | 1 |
| L | 8902 hCoV-19/USA/WI-WSLH-200312/2020\|EPI_ISL_578493\|202 | ···················· | ···················· | ···················· | ···················· | NNNNNNNNNNNNNNNNNNNN | NNNNNNNNNNNNNNNNNNNN | 1 |
| L | 8904 hCoV-19/USA/WI-WSLH-200314/2020\|EPI_ISL_578495\|202 | ···················· | ···················· | ···················· | ···················· | NNNNNNNNNNNNNNNNNNNN | NNNNNNNNNNNNNNNNNNNN | 1 |
| L | 8905 hCoV-19/USA/WI-WSLH-200315/2020\|EPI_ISL_578496\|202 | ···················· | ···················· | ···················· | ···················· | NNNNNNNNNNNNNNNNNNNN | NNNNNNNNNNNNNNNNNNNN | 1 |
| L | 8906 hCoV-19/USA/WI-WSLH-200316/2020\|EPI_ISL_578497\|202 | ···················· | ···················· | ···················· | ···················· | NNNNNNNNNNNNNNNNNNNN | NNNNNNNNNNNNNNNNNNNN | 1 |
| L | 8907 hCoV-19/USA/WI-WSLH-200317/2020\|EPI_ISL_578498\|202 | ···················· | ···················· | ···················· | ···················· | NNNNNNNNNNNNNNNNNNNN | NNNNNNNNNNNNNNNNNNNN | 1 |
| L | 8912 hCoV-19/USA/CA-CZB-10861/2020\|EPI_ISL_583103\|2020- | ···················· | ·A·················· | ···················· | ···················· | NNNNNNNNNNNNNNNNNNNN | NNNNNNNNNNNNNNNNNNNN | 1 |
| L | 8918 hCoV-19/USA/CA-CZB-10870/2020\|EPI_ISL_583110\|2020- | ···················· | ·A·················· | ···················· | ···················· | NNNN················ | ···················· | 1 |
| L | 8919 hCoV-19/USA/CA-CZB-10871/2020\|EPI_ISL_583111\|2020- | ···················· | ·A·················· | ···················· | ···················· | NNNN················ | ··NNNNNNNNNNNNN····· | 1 |
| L | 8925 hCoV-19/USA/CA-CZB-10879/2020\|EPI_ISL_583117\|2020- | ···················· | ·A·················· | ···················· | ···················· | NNNNNNNNNNNNNNNNNNNN | NNNNNNNNNNNNNNNNNNNN | 1 |
| L | 9030 hCoV-19/England/NOTT-11416F/2020\|EPI_ISL_577143\|20 | ···················· | ···················· | ···················· | ···················· | NNNNNNNNNNNNNNNNNNNN | NNNNNNNNNNNNNNNNNNNN | 1 |
| L | 9037 hCoV-19/England/NOTT-11427B/2020\|EPI_ISL_577153\|20 | ···················· | ···················· | ···················· | ···················· | NNNNNNNNNNNNNNNNNNNN | NNNNNNNNNNNNNNNNNNNN | 1 |
| L | 9040 hCoV-19/USA/WI-WSLH-200178/2020\|EPI_ISL_578404\|202 | ···················· | ···················· | ···················· | ···················· | NNNNNNNNNNNNNNNNNNNN | NNNNNNNNNNNNNNNNNNNN | 1 |
| L | 9042 hCoV-19/USA/WI-WSLH-200181/2020\|EPI_ISL_578406\|202 | ···················· | ···················· | ···················· | ···················· | NNNNNNNNNNNNNNNNNNNN | NNNNNNNNNNNNNNNNNNNN | 1 |
| L | 9045 hCoV-19/USA/WI-WSLH-200184/2020\|EPI_ISL_578409\|202 | ···················· | ···················· | ···················· | ···················· | NNNNNNNNNNNNNNNNNNNN | NNNNNNNNNNNNNNNNNNNN | 1 |
| L | 9046 hCoV-19/USA/WI-WSLH-200185/2020\|EPI_ISL_578410\|202 | ···················· | ···················· | ···················· | ···················· | NNNNNNNNNNNNNNNNNNNN | NNNNNNNNNNNNNNNNNNNN | 1 |
| L | 9047 hCoV-19/USA/WI-WSLH-200186/2020\|EPI_ISL_578411\|202 | ···················· | ···················· | ···················· | ···················· | NNNNNNNNNNNNNNNNNNNN | NNNNNNNNNNNNNNNNNNNN | 1 |
| L | 9048 hCoV-19/USA/WI-WSLH-200187/2020\|EPI_ISL_578412\|202 | ···················· | ···················· | ···················· | ···················· | NNNNNNNNNNNNNNNNNNNN | NNNNNNNNNNNNNNNNNNNN | 1 |
| L | 9049 hCoV-19/USA/WI-WSLH-200188/2020\|EPI_ISL_578413\|202 | ···················· | ···················· | ···················· | ···················· | NNNNNNNNNNNNNNNNNNNN | NNNNNNNNNNNNNNNNNNNN | 1 |
| L | 9050 hCoV-19/USA/WI-WSLH-200189/2020\|EPI_ISL_578414\|202 | ···················· | ···················· | ···················· | ···················· | NNNNNNNNNNNNNNNNNNNN | NNNNNNNNNNNNNNNNNNNN | 1 |
| L | 9051 hCoV-19/USA/WI-WSLH-200190/2020\|EPI_ISL_578415\|202 | ···················· | ···················· | ···················· | ···················· | NNNNNNNNNNNNNNNNNNNN | NNNNNNNNNNNNNNNNNNNN | 1 |
| L | 9057 hCoV-19/USA/WI-WSLH-200197/2020\|EPI_ISL_578421\|202 | ···················· | ···················· | ···················· | ···················· | NNNNNNNNNNNNNNNNNNNN | NNNNNNNNNNNNNNNNNNNN | 1 |
| L | 9058 hCoV-19/USA/WI-WSLH-200199/2020\|EPI_ISL_578422\|202 | ···················· | ···················· | ···················· | ···················· | NNNNNNNNNNNNNNNNNNNN | NNNNNNNNNNNNNNNNNNNN | 1 |
| L | 9059 hCoV-19/USA/WI-WSLH-200200/2020\|EPI_ISL_578423\|202 | ···················· | ···················· | ···················· | ···················· | NNNNNNNNNNNNNNNNNNNN | NNNNNNNNNNNNNNNNNNNN | 1 |
| L | 9060 hCoV-19/USA/WI-WSLH-200201/2020\|EPI_ISL_578424\|202 | ···················· | ···················· | ···················· | ···················· | NNNNNNNNNNNNNNNNNNNN | NNNNNNNNNNNNNNNNNNNN | 1 |
| L | 9061 hCoV-19/USA/WI-WSLH-200202/2020\|EPI_ISL_578425\|202 | ···················· | ···················· | ···················· | ···················· | NNNNNNNNNNNNNNNNNNNN | NNNNNNNNNNNNNNNNNNNN | 1 |
| L | 9062 hCoV-19/USA/WI-WSLH-200203/2020\|EPI_ISL_578426\|202 | ···················· | ···················· | ···················· | ···················· | NNNNNNNNNNNNNNNNNNNN | NNNNNNNNNNNNNNNNNNNN | 1 |
| L | 9063 hCoV-19/USA/WI-WSLH-200205/2020\|EPI_ISL_578427\|202 | ···················· | ···················· | ···················· | ···················· | NNNNNNNNNNNNNNNNNNNN | NNNNNNNNNNNNNNNNNNNN | 1 |
| L | 9064 hCoV-19/USA/WI-WSLH-200207/2020\|EPI_ISL_578428\|202 | ···················· | ···················· | ···················· | ···················· | NNNNNNNNNNNNNNNNNNNN | NNNNNNNNNNNNNNNNNNNN | 1 |
| L | 9065 hCoV-19/USA/WI-WSLH-200208/2020\|EPI_ISL_578429\|202 | ···················· | ···················· | ···················· | ···················· | NNNNNNNNNNNNNNNNNNNN | NNNNNNNNNNNNNNNNNNNN | 1 |
| L | 9066 hCoV-19/USA/WI-WSLH-200209/2020\|EPI_ISL_578430\|202 | ···················· | ···················· | ···················· | ···················· | NNNNNNNNNNNNNNNNNNNN | NNNNNNNNNNNNNNNNNNNN | 1 |
| L | 9067 hCoV-19/USA/WI-WSLH-200210/2020\|EPI_ISL_578431\|202 | ···················· | ···················· | ···················· | ···················· | NNNNNNNNNNNNNNNNNNNN | NNNNNNNNNNNNNNNNNNNN | 1 |
| L | 9068 hCoV-19/USA/WI-WSLH-200211/2020\|EPI_ISL_578432\|202 | ···················· | ···················· | ···················· | ···················· | NNNNNNNNNNNNNNNNNNNN | NNNNNNNNNNNNNNNNNNNN | 1 |

| | | | | | | | | |
|---|---|---|---|---|---|---|---|---|
| L | 9069 hCoV-19/USA/WI-WSLH-200212/2020\|EPI_ISL_578433\|202 | ···················· | ···················· | ···················· | ···················· | NNNNNNNNNNNNNNNNNNNN | NNNNNNNNNNNNNNNNNNNN | 1 |
| L | 9070 hCoV-19/USA/WI-WSLH-200213/2020\|EPI_ISL_578434\|202 | ···················· | ···················· | ···················· | ···················· | NNNNNNNNNNNNNNNNNNNN | NNNNNNNNNNNNNNNNNNNN | 1 |
| L | 9072 hCoV-19/USA/WI-WSLH-200215/2020\|EPI_ISL_578436\|202 | ···················· | ···················· | ···················· | ···················· | NNNNNNNNNNNNNNNNNNNN | NNNNNNNNNNNNNNNNNNNN | 1 |
| L | 9073 hCoV-19/USA/WI-WSLH-200217/2020\|EPI_ISL_578437\|202 | ···················· | ···················· | ···················· | ···················· | NNNNNNNNNNNNNNNNNNNN | NNNNNNNNNNNNNNNNNNNN | 1 |
| L | 9075 hCoV-19/USA/WI-WSLH-200219/2020\|EPI_ISL_578439\|202 | ···················· | ···················· | ···················· | ···················· | NNNNNNNNNNNNNNNNNNNN | NNNNNNNNNNNNNNNNNNNN | 1 |
| L | 9076 hCoV-19/USA/WI-WSLH-200220/2020\|EPI_ISL_578440\|202 | ···················· | ···················· | ···················· | ···················· | NNNNNNNNNNNNNNNNNNNN | NNNNNNNNNNNNNNNNNNNN | 1 |
| L | 9078 hCoV-19/USA/WI-WSLH-200222/2020\|EPI_ISL_578442\|202 | ···················· | ···················· | ···················· | ···················· | NNNNNNNNNNNNNNNNNNNN | NNNNNNNNNNNNNNNNNNNN | 1 |
| L | 9079 hCoV-19/USA/WI-WSLH-200223/2020\|EPI_ISL_578443\|202 | ···················· | ···················· | ···················· | ···················· | NNNNNNNNNNNNNNNNNNNN | NNNNNNNNNNNNNNNNNNNN | 1 |
| L | 9080 hCoV-19/USA/WI-WSLH-200224/2020\|EPI_ISL_578444\|202 | ···················· | ···················· | ···················· | ···················· | NNNNNNNNNNNNNNNNNNNN | NNNNNNNNNNNNNNNNNNNN | 1 |
| L | 9082 hCoV-19/USA/WI-WSLH-200226/2020\|EPI_ISL_578446\|202 | ···················· | ···················· | ···················· | ···················· | NNNNNNNNNNNNNNNNNNNN | NNNNNNNNNNNNNNNNNNNN | 1 |
| L | 9083 hCoV-19/USA/WI-WSLH-200227/2020\|EPI_ISL_578447\|202 | ···················· | ···················· | ···················· | ···················· | NNNNNNNNNNNNNNNNNNNN | NNNNNNNNNNNNNNNNNNNN | 1 |
| L | 9084 hCoV-19/USA/WI-WSLH-200228/2020\|EPI_ISL_578448\|202 | ···················· | ···················· | ···················· | ···················· | NNNNNNNNNNNNNNNNNNNN | NNNNNNNNNNNNNNNNNNNN | 1 |
| L | 9085 hCoV-19/USA/WI-WSLH-200229/2020\|EPI_ISL_578449\|202 | ···················· | ···················· | ···················· | ···················· | NNNNNNNNNNNNNNNNNNNN | NNNNNNNNNNNNNNNNNNNN | 1 |
| L | 9088 hCoV-19/USA/WI-WSLH-200232/2020\|EPI_ISL_578452\|202 | ···················· | ···················· | ···················· | ···················· | NNNNNNNNNNNNNNNNNNNN | NNNNNNNNNNNNNNNNNNNN | 1 |
| L | 9090 hCoV-19/USA/WI-WSLH-200234/2020\|EPI_ISL_578454\|202 | ···················· | ···················· | ···················· | ···················· | NNNNNNNNNNNNNNNNNNNN | NNNNNNNNNNNNNNNNNNNN | 1 |
| L | 9092 hCoV-19/USA/WI-WSLH-200236/2020\|EPI_ISL_578456\|202 | ···················· | ···················· | ···················· | ···················· | NNNNNNNNNNNNNNNNNNNN | NNNNNNNNNNNNNNNNNNNN | 1 |
| L | 9093 hCoV-19/USA/WI-WSLH-200237/2020\|EPI_ISL_578457\|202 | ···················· | ···················· | ···················· | ···················· | NNNNNNNNNNNNNNNNNNNN | NNNNNNNNNNNNNNNNNNNN | 1 |
| L | 9094 hCoV-19/USA/WI-WSLH-200238/2020\|EPI_ISL_578458\|202 | ···················· | ···················· | ···················· | ···················· | NNNNNNNNNNNNNNNNNNNN | NNNNNNNNNNNNNNNNNNNN | 1 |
| L | 9095 hCoV-19/USA/WI-WSLH-200239/2020\|EPI_ISL_578459\|202 | ···················· | ···················· | ···················· | ···················· | NNNNNNNNNNNNNNNNNNNN | NNNNNNNNNNNNNNNNNNNN | 1 |
| L | 9096 hCoV-19/USA/WI-WSLH-200240/2020\|EPI_ISL_578460\|202 | ···················· | ···················· | ···················· | ···················· | NNNNNNNNNNNNNNNNNNNN | NNNNNNNNNNNNNNNNNNNN | 1 |
| L | 9097 hCoV-19/USA/WI-WSLH-200241/2020\|EPI_ISL_578461\|202 | ···················· | ···················· | ···················· | ···················· | NNNNNNNNNNNNNNNNNNNN | NNNNNNNNNNNNNNNNNNNN | 1 |
| L | 9098 hCoV-19/USA/WI-WSLH-200242/2020\|EPI_ISL_578462\|202 | ···················· | ···················· | ···················· | ···················· | NNNNNNNNNNNNNNNNNNNN | NNNNNNNNNNNNNNNNNNNN | 1 |
| L | 9099 hCoV-19/USA/WI-WSLH-200244/2020\|EPI_ISL_578463\|202 | ···················· | ···················· | ···················· | ···················· | NNNNNNNNNNNNNNNNNNNN | NNNNNNNNNNNNNNNNNNNN | 1 |
| L | 9100 hCoV-19/USA/WI-WSLH-200246/2020\|EPI_ISL_578464\|202 | ···················· | ···················· | ···················· | ···················· | NNNNNNNNNNNNNNNNNNNN | NNNNNNNNNNNNNNNNNNNN | 1 |
| L | 9101 hCoV-19/USA/WI-WSLH-200247/2020\|EPI_ISL_578465\|202 | ···················· | ···················· | ···················· | ···················· | NNNNNNNNNNNNNNNNNNNN | NNNNNNNNNNNNNNNNNNNN | 1 |
| L | 9102 hCoV-19/USA/WI-WSLH-200250/2020\|EPI_ISL_578466\|202 | ···················· | ···················· | ···················· | ···················· | NNNNNNNNNNNNNNNNNNNN | NNNNNNNNNNNNNNNNNNNN | 1 |
| L | 9103 hCoV-19/USA/WI-WSLH-200251/2020\|EPI_ISL_578467\|202 | ···················· | ···················· | ···················· | ···················· | NNNNNNNNNNNNNNNNNNNN | NNNNNNNNNNNNNNNNNNNN | 1 |
| L | 9104 hCoV-19/USA/WI-WSLH-200253/2020\|EPI_ISL_578468\|202 | ···················· | ···················· | ···················· | ···················· | NNNNNNNNNNNNNNNNNNNN | NNNNNNNNNNNNNNNNNNNN | 1 |
| L | 9105 hCoV-19/USA/WI-WSLH-200256/2020\|EPI_ISL_578469\|202 | ···················· | ···················· | ···················· | ···················· | NNNNNNNNNNNNNNNNNNNN | NNNNNNNNNNNNNNNNNNNN | 1 |
| L | 9106 hCoV-19/USA/WI-WSLH-200263/2020\|EPI_ISL_578473\|202 | ···················· | ···················· | ···················· | ···················· | NNNNNNNNNNNNNNNNNNNN | NNNNNNNNNNNNNNNNNNNN | 1 |
| L | 9107 hCoV-19/USA/WI-WSLH-200287/2020\|EPI_ISL_578474\|202 | ···················· | ···················· | ···················· | ···················· | NNNNNNNNNNNNNNNNNNNN | NNNNNNNNNNNNNNNNNNNN | 1 |
| L | 9108 hCoV-19/USA/WI-WSLH-200288/2020\|EPI_ISL_578475\|202 | ···················· | ···················· | ···················· | ···················· | NNNNNNNNNNNNNNNNNNNN | NNNNNNNNNNNNNNNNNNNN | 1 |
| L | 9109 hCoV-19/USA/WI-WSLH-200289/2020\|EPI_ISL_578476\|202 | ···················· | ···················· | ···················· | ···················· | NNNNNNNNNNNNNNNNNNNN | NNNNNNNNNNNNNNNNNNNN | 1 |
| L | 9110 hCoV-19/USA/WI-WSLH-200291/2020\|EPI_ISL_578477\|202 | ···················· | ···················· | ···················· | ···················· | NNNNNNNNNNNNNNNNNNNN | NNNNNNNNNNNNNNNNNNNN | 1 |
| L | 9111 hCoV-19/USA/WI-WSLH-200292/2020\|EPI_ISL_578478\|202 | ···················· | ···················· | ···················· | ···················· | NNNNNNNNNNNNNNNNNNNN | NNNNNNNNNNNNNNNNNNNN | 1 |
| L | 9112 hCoV-19/USA/WI-WSLH-200293/2020\|EPI_ISL_578479\|202 | ···················· | ···················· | ···················· | ···················· | NNNNNNNNNNNNNNNNNNNN | NNNNNNNNNNNNNNNNNNNN | 1 |
| L | 9113 hCoV-19/USA/WI-WSLH-200295/2020\|EPI_ISL_578480\|202 | ···················· | ···················· | ···················· | ···················· | NNNNNNNNNNNNNNNNNNNN | NNNNNNNNNNNNNNNNNNNN | 1 |
| L | 9114 hCoV-19/USA/WI-WSLH-200297/2020\|EPI_ISL_578481\|202 | ···················· | ···················· | ···················· | ···················· | NNNNNNNNNNNNNNNNNNNN | NNNNNNNNNNNNNNNNNNNN | 1 |
| L | 9115 hCoV-19/USA/WI-WSLH-200298/2020\|EPI_ISL_578482\|202 | ···················· | ···················· | ···················· | ···················· | NNNNNNNNNNNNNNNNNNNN | NNNNNNNNNNNNNNNNNNNN | 1 |
| L | 9116 hCoV-19/USA/WI-WSLH-200299/2020\|EPI_ISL_578483\|202 | ···················· | ···················· | ···················· | ···················· | NNNNNNNNNNNNNNNNNNNN | NNNNNNNNNNNNNNNNNNNN | 1 |
| L | 9117 hCoV-19/USA/WI-WSLH-200300/2020\|EPI_ISL_578484\|202 | ···················· | ···················· | ···················· | ···················· | NNNNNNNNNNNNNNNNNNNN | NNNNNNNNNNNNNNNNNNNN | 1 |
| L | 9118 hCoV-19/USA/WI-WSLH-200301/2020\|EPI_ISL_578485\|202 | ···················· | ···················· | ···················· | ···················· | NNNNNNNNNNNNNNNNNNNN | NNNNNNNNNNNNNNNNNNNN | 1 |
| L | 9119 hCoV-19/USA/WI-WSLH-200302/2020\|EPI_ISL_578486\|202 | ···················· | ···················· | ···················· | ···················· | NNNNNNNNNNNNNNNNNNNN | NNNNNNNNNNNNNNNNNNNN | 1 |
| L | 9120 hCoV-19/USA/WI-WSLH-200303/2020\|EPI_ISL_578487\|202 | ···················· | ···················· | ···················· | ···················· | NNNNNNNNNNNNNNNNNNNN | NNNNNNNNNNNNNNNNNNNN | 1 |
| L | 9121 hCoV-19/USA/WI-WSLH-200306/2020\|EPI_ISL_578488\|202 | ···················· | ···················· | ···················· | ···················· | NNNNNNNNNNNNNNNNNNNN | NNNNNNNNNNNNNNNNNNNN | 1 |
| L | 9122 hCoV-19/USA/WI-WSLH-200307/2020\|EPI_ISL_578489\|202 | ···················· | ···················· | ···················· | ···················· | NNNNNNNNNNNNNNNNNNNN | NNNNNNNNNNNNNNNNNNNN | 1 |
| L | 9301 hCoV-19/USA/WI-WSLH-200261/2020\|EPI_ISL_578471\|202 | ···················· | ···················· | ···················· | ···················· | NNNNNNNNNNNNNNNNNNNN | NNNNNNNNNNNNNNNNNNNN | 1 |
| L | 9302 hCoV-19/USA/WI-WSLH-200262/2020\|EPI_ISL_578472\|202 | ···················· | ···················· | ···················· | ···················· | NNNNNNNNNNNNNNNNNNNN | NNNNNNNNNNNNNNNNNNNN | 1 |
| L | 9304 hCoV-19/USA/WI-WSLH-200320/2020\|EPI_ISL_578501\|202 | ···················· | ···················· | ···················· | ···················· | NNNNNNNNNNNNNNNNNNNN | NNNNNNNNNNNNNNNNNNNN | 1 |
| L | 9308 hCoV-19/USA/WI-WSLH-200324/2020\|EPI_ISL_578505\|202 | ···················· | ···················· | ···················· | ···················· | NNNNNNNNNNNNNNNNNNNN | NNNNNNNNNNNNNNNNNNNN | 1 |
| L | 9309 hCoV-19/USA/WI-WSLH-200325/2020\|EPI_ISL_578506\|202 | ···················· | ···················· | ···················· | ···················· | NNNNNNNNNNNNNNNNNNNN | NNNNNNNNNNNNNNNNNNNN | 1 |
| L | 9310 hCoV-19/USA/WI-WSLH-200326/2020\|EPI_ISL_578507\|202 | ···················· | ···················· | ···················· | ···················· | NNNNNNNNNNNNNNNNNNNN | NNNNNNNNNNNNNNNNNNNN | 1 |
| L | 9311 hCoV-19/USA/WI-WSLH-200327/2020\|EPI_ISL_578508\|202 | ···················· | ···················· | ···················· | ···················· | NNNNNNNNNNNNNNNNNNNN | NNNNNNNNNNNNNNNNNNNN | 1 |
| L | 9314 hCoV-19/USA/WI-WSLH-200330/2020\|EPI_ISL_578511\|202 | ···················· | ···················· | ···················· | ···················· | NNNNNNNNNNNNNNNNNNNN | NNNNNNNNNNNNNNNNNNNN | 1 |
| L | 9316 hCoV-19/USA/WI-WSLH-200332/2020\|EPI_ISL_578513\|202 | ···················· | ···················· | ···················· | ···················· | NNNNNNNNNNNNNNNNNNNN | NNNNNNNNNNNNNNNNNNNN | 1 |
| L | 9317 hCoV-19/USA/WI-WSLH-200339/2020\|EPI_ISL_578514\|202 | ···················· | ···················· | ···················· | ···················· | NNNNNNNNNNNNNNNNNNNN | NNNNNNNNNNNNNNNNNNNN | 1 |
| L | 9319 hCoV-19/USA/WI-WSLH-200341/2020\|EPI_ISL_578516\|202 | ···················· | ···················· | ···················· | ···················· | NNNNNNNNNNNNNNNNNNNN | NNNNNNNNNNNNNNNNNNNN | 1 |
| L | 9320 hCoV-19/USA/WI-WSLH-200342/2020\|EPI_ISL_578517\|202 | ···················· | ···················· | ···················· | ···················· | NNNNNNNNNNNNNNNNNNNN | NNNNNNNNNNNNNNNNNNNN | 1 |
| L | 9321 hCoV-19/USA/WI-WSLH-200343/2020\|EPI_ISL_578518\|202 | ···················· | ···················· | ···················· | ···················· | NNNNNNNNNNNNNNNNNNNN | NNNNNNNNNNNNNNNNNNNN | 1 |
| L | 9322 hCoV-19/USA/WI-WSLH-200345/2020\|EPI_ISL_578519\|202 | ···················· | ···················· | ···················· | ···················· | NNNNNNNNNNNNNNNNNNNN | NNNNNNNNNNNNNNNNNNNN | 1 |
| L | 9325 hCoV-19/USA/WI-WSLH-200351/2020\|EPI_ISL_578522\|202 | ···················· | ···················· | ···················· | ···················· | NNNNNNNNNNNNNNNNNNNN | NNNNNNNNNNNNNNNNNNNN | 1 |
| L | 9326 hCoV-19/USA/WI-WSLH-200352/2020\|EPI_ISL_578523\|202 | ···················· | ···················· | ···················· | ···················· | NNNNNNNNNNNNNNNNNNNN | NNNNNNNNNNNNNNNNNNNN | 1 |
| L | 9327 hCoV-19/USA/WI-WSLH-200353/2020\|EPI_ISL_578524\|202 | ···················· | ···················· | ···················· | ···················· | NNNNNNNNNNNNNNNNNNNN | NNNNNNNNNNNNNNNNNNNN | 1 |
| L | 9456 hCoV-19/USA/CA-CZB-10836/2020\|EPI_ISL_583083\|2020- | ···················· | ···················· | ···················· | ···················· | NNNNNNNNNNNN·NNNNNNN | ···················· | 1 |
| M | 5 hCoV-19/Austria/CeMM0812/2020\|EPI_ISL_583714\|2020- | ···················· | ···················· | ···················· | ···················· | NNNNNNNNNNNNNNNNNNNN | NNNNNNNNNNNNNNNNNNNN | 1 |
| M | 33 hCoV-19/Austria/CeMM0891/2020\|EPI_ISL_583744\|2020- | ···················· | ···················· | ···················· | ···················· | NNNNNNNNNNNNNNNNNNNN | NNNNNNNNNNNNNNNNNNNN | 1 |
| M | 45 hCoV-19/Austria/CeMM0903/2020\|EPI_ISL_583756\|2020- | ···················· | ···················· | ···················· | ···················· | NNNNNNNNNNNNNNNNNNNN | NNNNNNNNNNNNNNNNNNNN | 1 |
| M | 153 hCoV-19/Australia/VIC15006/2020\|EPI_ISL_593100\|202 | ···················· | ···················· | ···················· | ···················· | NNNNNNNNNNNNNNNNNNNN | NNNNNNNNNNNNNNNNNNNN | 1 |
| M | 154 hCoV-19/Australia/VIC15011/2020\|EPI_ISL_593101\|202 | ···················· | ···················· | ···················· | ···················· | NNNNNNNNNNNNNNNNNNNN | NNNNNNNNNNNNNNNNNNNN | 1 |
| M | 157 hCoV-19/Australia/VIC15017/2020\|EPI_ISL_593104\|202 | ···················· | ···················· | ···················· | ···················· | NNNNNNNNNNNNNNNNNNNN | NNNNNNNNNNNNNNNNNNNN | 1 |
| M | 159 hCoV-19/Australia/VIC15019/2020\|EPI_ISL_593106\|202 | ···················· | ···················· | ···················· | ···················· | NNNN················ | ···················· | 1 |
| M | 268 hCoV-19/Wales/PHWC-16F6DF/2020\|EPI_ISL_586007\|2020 | ···················· | ···················· | ···················· | ···················· | ···················· | NNNNN··············· | 1 |
| M | 277 hCoV-19/Wales/PHWC-16F7BE/2020\|EPI_ISL_586019\|2020 | ···················· | ···················· | ···················· | ···················· | NNNNNNNNNNNNNNNNNNNN | ···················· | 1 |
| M | 337 hCoV-19/England/QEUH-A1DCE4/2020\|EPI_ISL_587355\|20 | ···················· | ···················· | ···················· | ···················· | NNNNNNNNNNNNNNNNNNNN | NNNNNNNNNNNNNNNNNNNN | 1 |
| M | 639 hCoV-19/Australia/VIC15028/2020\|EPI_ISL_593111\|202 | ···················· | ···················· | ···················· | ···················· | NNNNNNNNNNNNNNNNNNNN | NNNNNNNNNNNNNNNNNNNN | 1 |
| M | 643 hCoV-19/Australia/VIC15032/2020\|EPI_ISL_593115\|202 | ···················· | ···················· | ···················· | ···················· | ·NNNNNNNNNNNNNNNNNNN | ···················· | 1 |
| M | 655 hCoV-19/Australia/VIC15073/2020\|EPI_ISL_593127\|202 | ···················· | ···················· | ···················· | ···················· | ······R···W·NNNNNNN | ···················· | 1 |
| M | 656 hCoV-19/Australia/VIC15076/2020\|EPI_ISL_593128\|202 | ···················· | ···················· | ···················· | ···················· | ···················· | NNNNNNNNN··········· | 1 |
| M | 659 hCoV-19/Australia/VIC15087/2020\|EPI_ISL_593131\|202 | ···················· | ···················· | ···················· | ···················· | NNNNNNNNNNNN········ | ···················· | 1 |
| M | 664 hCoV-19/Australia/VIC15097/2020\|EPI_ISL_593136\|202 | ···················· | ···················· | ···················· | ···················· | NNNNNNNNNNNNNNNNNNNN | ···················· | 1 |

| | | | | | | | | |
|---|---|---|---|---|---|---|---|---|
| M | 666 hCoV-19/Australia/VIC15105/2020\|EPI_ISL_593138\|202 | ···················· | ···················· | ···················· | ···················· | NNNN················ | ···················· | 1 |
| M | 673 hCoV-19/Australia/VIC15118/2020\|EPI_ISL_593145\|202 | ···················· | ···················· | ···················· | ···················· | NNNNNNNNNNNNNNNNNNNN | ···················· | 1 |
| M | 682 hCoV-19/Australia/VIC15139/2020\|EPI_ISL_593154\|202 | ···················· | ···················· | ···················· | NNNNNNNNNNNNNNNNNNNN | ···················· | ···················· | 1 |
| M | 685 hCoV-19/Australia/VIC15158/2020\|EPI_ISL_593164\|202 | ···················· | ···················· | ···················· | ···················· | NNNNNNNNNNNNNNNN···· | ···················· | 1 |
| M | 702 hCoV-19/Australia/VIC15213/2020\|EPI_ISL_593192\|202 | ···················· | ···················· | ···················· | ···················· | NNNNNNNNNN·········· | ···················· | 1 |
| M | 705 hCoV-19/Australia/VIC15216/2020\|EPI_ISL_593195\|202 | ···················· | ···················· | ···················· | ···················· | NNNNNNNNNNNNNNNNNNNN | NNNNNNNNNNNNNNNNNNNN | 1 |
| M | 738 hCoV-19/England/BIRM-6335B/2020\|EPI_ISL_594484\|202 | ···················· | ···················· | ···················· | ···················· | NNNNNNNNNNNNNNNNNNNN | NNNNNNNNNNNNNNNNNNNN | 1 |
| M | 739 hCoV-19/England/BIRM-6427D/2020\|EPI_ISL_594485\|202 | ···················· | ···················· | ···················· | ···················· | NNNNNNNNNNNNNNNNNNNN | NNNNNNNNNNNNNNNNNNNN | 1 |
| M | 741 hCoV-19/England/BIRM-6444A/2020\|EPI_ISL_594494\|202 | ···················· | ·A·················· | ···················· | ···················· | NNNNNNNNNNNNNNNNNNNN | NNNNNNNNNNNNNNNNNNNN | 1 |
| M | 835 hCoV-19/England/QEUH-A029AF/2020\|EPI_ISL_584939\|20 | ···················· | ···················· | ···················· | ···················· | NNNNNNNNNNNNNNNNNNNN | NNNNNNNNNNNNNNNNNNNN | 1 |
| M | 836 hCoV-19/England/QEUH-A02ACA/2020\|EPI_ISL_584945\|20 | ···················· | ···················· | ···················· | ···················· | NNNNNNNNNNNNNNNNNNNN | NNNNNNNNNNNNNNNNNNNN | 1 |
| M | 837 hCoV-19/England/QEUH-A02B12/2020\|EPI_ISL_584947\|20 | ···················· | ···················· | ···················· | ···················· | NNNNNNNNNNNNNNNNNNNN | NNNNNNNNNNNNNNNNNNNN | 1 |
| M | 838 hCoV-19/England/QEUH-A0C26E/2020\|EPI_ISL_584968\|20 | ···················· | ···················· | ···················· | ···················· | NNNNNNNNNNNNNNNNNNNN | NNNNNNNNNNNNNNNNNNNN | 1 |
| M | 939 hCoV-19/Australia/VIC15140/2020\|EPI_ISL_593155\|202 | ···················· | ···················· | ···················· | ···················· | NNNNNNNNNNNNNNNNNNNN | NNNNNNNNNNNNNNNNNNNN | 1 |
| M | 947 hCoV-19/Australia/VIC15164/2020\|EPI_ISL_593167\|202 | ···················· | ···················· | ···················· | ···················· | NNNNNNNNNNNNNNNNNNNN | NNNNNNNNNNNNNNNNNNNN | 1 |
| M | 950 hCoV-19/Australia/VIC15171/2020\|EPI_ISL_593170\|202 | ···················· | ···················· | ···················· | ···················· | NNNNNNNNNNNNNNNNNNNN | NNNNNNNNNNNNNNNNNNNN | 1 |
| M | 969 hCoV-19/USA/FL-BPHL-1623/2020\|EPI_ISL_594313\|2020- | ···················· | ···················· | ···················· | ···················· | NNNNNNNNNNNNNNNNN··· | ···················· | 1 |
| M | 972 hCoV-19/England/BIRM-6429B/2020\|EPI_ISL_594486\|202 | ···················· | ·A·················· | ···················· | ···················· | NNNNNNNNNNNNNNNNNNNN | NNNNNNNNNNNNNNNNNNNN | 1 |
| M | 974 hCoV-19/England/BIRM-6436B/2020\|EPI_ISL_594489\|202 | ···················· | ·A·················· | ···················· | ···················· | NNNNNNNNNNNNNNNNNNNN | NNNNNNNNNNNNNNNNNNNN | 1 |
| M | 975 hCoV-19/England/BIRM-6437A/2020\|EPI_ISL_594490\|202 | ···················· | ·A·················· | ···················· | ···················· | NNNNNNNNNNNNNNNNNNNN | NNNNNNNNNNNNNNNNNNNN | 1 |
| M | 976 hCoV-19/England/BIRM-645ED/2020\|EPI_ISL_594497\|202 | ···················· | ···················· | ···················· | ···················· | NNNNNNNNNNNNNNNNNNNN | NNNNNNNNNNNNNNNNNNNN | 1 |
| M | 988 hCoV-19/Wales/PHWC-47EF48/2020\|EPI_ISL_595615\|2020 | ···················· | ·A·················· | ···················· | ···················· | NNNNNNNNNNNNNNNNNNNN | NNNNNNNNNNNNNNNNNNNN | 1 |
| M | 1029 hCoV-19/England/QEUH-A02602/2020\|EPI_ISL_584915\|20 | ···················· | ···················· | ···················· | ···················· | NNNNNNNNNNNNNNNNNNNN | NNNNNNNNNNNNNNNNNNNN | 1 |
| M | 1261 hCoV-19/Australia/VIC14957/2020\|EPI_ISL_593080\|202 | ···················· | ···················· | ···················· | ···················· | NNNNNNNNNNNNNNNNNNNN | ···················· | 1 |
| M | 1262 hCoV-19/Australia/VIC14958/2020\|EPI_ISL_593081\|202 | ···················· | ···················· | ···················· | ···················· | NNNNNNNNNNNN········ | ···················· | 1 |
| M | 1270 hCoV-19/Australia/VIC14978/2020\|EPI_ISL_593089\|202 | ···················· | ···················· | ···················· | ···················· | NNNNNNNNNNNNNN······ | ···················· | 1 |
| M | 1274 hCoV-19/Australia/VIC14987/2020\|EPI_ISL_593093\|202 | ···················· | ···················· | ···················· | ···················· | NNNNNNNNNNNNNNNNNNNN | NNNNNNNNNNNNNNNNNNNN | 1 |
| M | 1276 hCoV-19/Australia/VIC14990/2020\|EPI_ISL_593095\|202 | ···················· | ···················· | ···················· | ···················· | NNNNNNNNNNNNNNNNNNNN | NNNNNNNNNNNNNNNNNNNN | 1 |
| M | 1278 hCoV-19/Australia/VIC14995/2020\|EPI_ISL_593097\|202 | ···················· | ···················· | ···················· | ···················· | NNNNNNNNNNNNNNNNNNNN | ···················· | 1 |
| M | 1279 hCoV-19/Australia/VIC14997/2020\|EPI_ISL_593098\|202 | ···················· | ···················· | ···················· | ···················· | NNNNNNNNNNNNNNNNNNNN | NNNNNNNNNNNNNNNNNNNN | 1 |
| M | 1289 hCoV-19/Austria/CeMM0745/2020\|EPI_ISL_583682\|2020- | ···················· | ···················· | ···················· | ···················· | NNNNNNNNNNNNNNNNNNNN | NNNNNNNNNNNNNNNNNNNN | 1 |
| M | 1397 hCoV-19/Australia/VIC14794/2020\|EPI_ISL_593001\|202 | ···················· | ···················· | ···················· | ···················· | NNNNNNNNNNNNNNNNNNNN | ···················· | 1 |
| M | 1398 hCoV-19/Australia/VIC14797/2020\|EPI_ISL_593002\|202 | ···················· | ···················· | ···················· | ···················· | NNNNNNNNNNNNNNNNNNNN | NNNNNNNNNNNNNNNNNNNN | 1 |
| M | 1400 hCoV-19/Australia/VIC14804/2020\|EPI_ISL_593004\|202 | ···················· | ···················· | ··K················· | ···················· | NNNNNNNNNNNNNNNNNNNN | NNNNNNNNNNNNNNNNNN·· | 1 |
| M | 1401 hCoV-19/Australia/VIC14806/2020\|EPI_ISL_593005\|202 | ···················· | ···················· | ···················· | ···················· | NNNNNNNNNNNN········ | ···················· | 1 |
| M | 1402 hCoV-19/Australia/VIC14807/2020\|EPI_ISL_593006\|202 | ···················· | ···················· | ···················· | ···················· | NNNNNNNNNNNNNNNNNNNN | NNNN················ | 1 |
| M | 1403 hCoV-19/Australia/VIC14813/2020\|EPI_ISL_593007\|202 | ···················· | ···················· | ···················· | ···················· | NNNNNNNNNNNN········ | ···················· | 1 |
| M | 1411 hCoV-19/Australia/VIC14826/2020\|EPI_ISL_593015\|202 | ···················· | ···················· | ···················· | ···················· | NNNNNNNNNNNNNNNNNNNN | ···················· | 1 |
| M | 1413 hCoV-19/Australia/VIC14829/2020\|EPI_ISL_593017\|202 | ···················· | ···················· | ···················· | ···················· | NNNNNNNNNNNN········ | ···················· | 1 |
| M | 1414 hCoV-19/Australia/VIC14832/2020\|EPI_ISL_593018\|202 | ···················· | ···················· | ···················· | ···················· | NNNNNNNNNNNNNNNNNNNN | NNNNNNNNNNNNNNNNNNNN | 1 |
| M | 1415 hCoV-19/Australia/VIC14835/2020\|EPI_ISL_593019\|202 | ···················· | ···················· | ···················· | ···················· | NNNNNNNNNNNNNNNNNNNN | NNNNNNNNNNNNNNNNNNNN | 1 |
| M | 1416 hCoV-19/Australia/VIC14836/2020\|EPI_ISL_593020\|202 | ···················· | ···················· | ···················· | ···················· | NNNNNNNNNNNNNNNNN··· | ···················· | 1 |
| M | 1417 hCoV-19/Australia/VIC14837/2020\|EPI_ISL_593021\|202 | ···················· | ···················· | ···················· | ···················· | NNNNNNNNNNNNNNNNNNNN | ······MM············ | 1 |
| M | 1418 hCoV-19/Australia/VIC14838/2020\|EPI_ISL_593022\|202 | ···················· | ···················· | ···················· | ···················· | NNNNNNNNNNNNNNNNNNNN | ···················· | 1 |
| M | 1422 hCoV-19/Australia/VIC14843/2020\|EPI_ISL_593026\|202 | ···················· | ···················· | ···················· | ···················· | NNNNNNNNNNNNNNNNNNNN | ···················· | 1 |
| M | 1426 hCoV-19/Australia/VIC14849/2020\|EPI_ISL_593030\|202 | ···················· | ···················· | ···················· | ···················· | ·······NN··········· | NNNNNNN············· | 1 |
| M | 1429 hCoV-19/Australia/VIC14861/2020\|EPI_ISL_593033\|202 | ···················· | ···················· | ···················· | ···················· | NNNNNNNNNNNNNNNNNNNN | ···················· | 1 |
| M | 1431 hCoV-19/Australia/VIC14877/2020\|EPI_ISL_593041\|202 | ···················· | ···················· | ···················· | ···················· | NNNNNNNNNNNNNNNNNNNN | NNNNNNNNNNNNNNNNNNNN | 1 |
| M | 1434 hCoV-19/Australia/VIC14885/2020\|EPI_ISL_593044\|202 | ···················· | ···················· | ···················· | ···················· | NNNNNNNNNNNNNNNN···· | ···················· | 1 |
| M | 1440 hCoV-19/Australia/VIC14923/2020\|EPI_ISL_593061\|202 | ···················· | ···················· | ···················· | ···················· | NNNNNNNNNNNNNNNNNNNN | NNNNNNNNNNNNNNNNNNNN | 1 |
| M | 1442 hCoV-19/Australia/VIC14926/2020\|EPI_ISL_593063\|202 | ···················· | ···················· | ···················· | ···················· | NNNNNNNNNNNNNNNNNNNN | NNNNNNNNNNNNNNNNNN·· | 1 |
| M | 1447 hCoV-19/Australia/VIC14934/2020\|EPI_ISL_593068\|202 | ···················· | ···················· | ···················· | ···················· | NNNNNNNNNNNNNNNNNNNN | ···················· | 1 |
| M | 1448 hCoV-19/Australia/VIC14935/2020\|EPI_ISL_593069\|202 | ···················· | ···················· | ···················· | ···················· | NNNNNNNNNNNNNNNNNNNN | NNNNNNNNNNNNNNNNNNNN | 1 |
| M | 1449 hCoV-19/Australia/VIC14937/2020\|EPI_ISL_593070\|202 | ···················· | ···················· | ···················· | ···················· | NNNNNNNNNNNNNNNNNNNN | ···················· | 1 |
| M | 1450 hCoV-19/Australia/VIC14939/2020\|EPI_ISL_593071\|202 | ···················· | ···················· | ···················· | ···················· | NNNNNNNNNNNNNNNNNNNN | NNNNNNNNNNNNNNN····· | 1 |
| M | 1451 hCoV-19/Australia/VIC14942/2020\|EPI_ISL_593072\|202 | ···················· | ···················· | ···················· | ···················· | NNNNNNNNNNNNNNNNNNNN | NNNNNNNNNNNNNNNNNNNN | 1 |
| M | 1456 hCoV-19/Australia/VIC14953/2020\|EPI_ISL_593077\|202 | ···················· | ···················· | ···················· | NNNNNNNNNNNNNNNNNNNN | ···················· | ···················· | 1 |
| M | 1460 hCoV-19/USA/FL-BPHL-1647/2020\|EPI_ISL_594336\|2020- | ···················· | ···················· | ···················· | ···················· | NNNNNNNNNNNNNNNNNNNN | ···················· | 1 |
| M | 1467 hCoV-19/USA/FL-BPHL-1656/2020\|EPI_ISL_594345\|2020- | ···················· | ···················· | ···················· | ···················· | NNNNNNNNNNNNNNNNNNNN | ···················· | 1 |
| M | 1473 hCoV-19/USA/FL-BPHL-1663/2020\|EPI_ISL_594352\|2020- | ···················· | ···················· | ···················· | ···················· | NNNN················ | ···················· | 1 |
| M | 1482 hCoV-19/USA/FL-BPHL-1675/2020\|EPI_ISL_594364\|2020- | ···················· | ········A··········· | ···················· | ···················· | NNNNNNNNNNNNNNNNNNNN | ···················· | 1 |
| M | 1484 hCoV-19/USA/FL-BPHL-1682/2020\|EPI_ISL_594371\|2020- | ···················· | ···················· | ···················· | ···················· | NNNNNNNNNNNNNNNNNNNN | ···················· | 1 |
| M | 1738 hCoV-19/Australia/VIC12461/2020\|EPI_ISL_591996\|202 | ···················· | ···················· | ···················· | ···················· | NNNN················ | ···················· | 1 |
| M | 1741 hCoV-19/Australia/VIC12468/2020\|EPI_ISL_591999\|202 | ···················· | ···················· | ···················· | ···················· | NNNNNNNNNNNNNNNNNNNN | NNNNNNNNNNNNNNN····· | 1 |
| M | 1743 hCoV-19/Australia/VIC14865/2020\|EPI_ISL_593035\|202 | ···················· | ···················· | ···················· | ···················· | NNNNNNNNNNNNNNNNNNNN | NNNNNNNNNNNNNNNNNNNN | 1 |
| M | 1747 hCoV-19/Australia/VIC14873/2020\|EPI_ISL_593039\|202 | ···················· | ···················· | ···················· | ···················· | NNNNNNNNNNNNNNNNNNNN | NNNNNNNNNNNNNNNNNNNN | 1 |
| M | 1752 hCoV-19/Australia/VIC14894/2020\|EPI_ISL_593049\|202 | ···················· | ···················· | ···················· | ···················· | NNNNNNNNNNNNNNNNNNNN | NNNNNNNNNNNNNNNNNNNN | 1 |
| M | 1754 hCoV-19/Australia/VIC14897/2020\|EPI_ISL_593051\|202 | ···················· | ···················· | ···················· | ···················· | NNNNNNNNNNNNNNNNNNNN | NNNNNNNNNNNNNNNNNNNN | 1 |
| M | 1755 hCoV-19/Australia/VIC14900/2020\|EPI_ISL_593052\|202 | ···················· | ···················· | ···················· | ···················· | NNNNNNNNNNNNNNNNNNNN | NNNNNNNNNNNNNNNNNNNN | 1 |
| M | 1756 hCoV-19/Australia/VIC14901/2020\|EPI_ISL_593053\|202 | ···················· | ···················· | ···················· | ···················· | NNNNNNNNNNNNNNNNNNNN | NNNNNNNNN··········M | 1 |
| M | 1758 hCoV-19/Australia/VIC14907/2020\|EPI_ISL_593055\|202 | ···················· | ···················· | ···················· | ···················· | NNNNNNNNNNNNNNNNNNNN | NNNNNNNNNNNNNNNNNNNN | 1 |
| M | 1766 hCoV-19/USA/FL-BPHL-1696/2020\|EPI_ISL_594385\|2020- | ···················· | ···················· | ···················· | ···················· | NNNNNNNNNNNNNNNNN··· | ···················· | 1 |
| M | 1773 hCoV-19/Wales/PHWC-47E801/2020\|EPI_ISL_595507\|2020 | ···················· | ···················· | ···················· | ···················· | ···················· | NNNNN··············· | 1 |
| M | 1780 hCoV-19/Wales/PHWC-47E8C5/2020\|EPI_ISL_595517\|2020 | ···················· | ···················· | ···················· | ···················· | NNNNNNNNNNNNNNNNNNNN | NNNNNNNNNNNNNNNNNNNN | 1 |
| M | 2004 hCoV-19/USA/MI-UM-10036118316/2020\|EPI_ISL_590714\| | ···················· | ···················· | ···················· | ···················· | NNNNNNNNNNNNNNNNNNNN | ···················· | 1 |
| M | 2010 hCoV-19/USA/MI-UM-10036196865/2020\|EPI_ISL_590740\| | ···················· | ···················· | ···················· | ···················· | NNNNNNNNNNNNNNNNNNNN | NNNNNNNNNNNNNNNNNNNN | 1 |
| M | 2021 hCoV-19/USA/MI-UM-MH27541/2020\|EPI_ISL_590751\|2020 | ···················· | ···················· | ···················· | ···················· | NNNNNNNNNNNNNNNNNNNN | NNNNNNNNNNNNNNNNNNNN | 1 |
| M | 2033 hCoV-19/Australia/VIC12420/2020\|EPI_ISL_591984\|202 | ···················· | ···················· | ···················· | ···················· | NNNNNNNNNNNNNNNNNNNN | NNNNNNNNNNNNNNNNNNNN | 1 |
| M | 2036 hCoV-19/Australia/VIC12428/2020\|EPI_ISL_591987\|202 | ···················· | ···················· | ···················· | ···················· | NNNNNNNNNNNNNNNNNNNN | ···················· | 1 |
| M | 2042 hCoV-19/USA/FL-BPHL-1601/2020\|EPI_ISL_594291\|2020- | ···················· | ···················· | ···················· | ···················· | NNNNNNNNNNNNNNNNNNNN | N··················· | 1 |

M 2045 hCoV-19/USA/FL-BPHL-1605/2020|EPI_ISL_594295|2020- ···················· ···················· ···················· ···················· NNNNNNNNNNNNNNNNNN·· ···················· 1
M 2080 hCoV-19/England/NORW-EF91B/2020|EPI_ISL_584717|202 ···················· ···················· ···················· ···················· NNNNNNNNNNNNNNNNNNNN NNNNNNNNNNNNNNNNNNNN 1
M 2081 hCoV-19/England/NORW-F0452/2020|EPI_ISL_584729|202 ···················· ···················· ···················· ···················· NNNNNNNNNNNNNNNNNNNN NNNNNNNNNNNNNNNNNNNN 1
M 2083 hCoV-19/England/NORW-F072C/2020|EPI_ISL_584738|202 ···················· ···················· ···················· ···················· NNNNNNNNNNNNNNNNNNNN NNNNNNNNNNNNNNNNNNNN 1
M 2084 hCoV-19/England/NORW-F0A23/2020|EPI_ISL_584746|202 ···················· ·A·················· ···················· ···················· NNNNNNNNNNNNNNNNNNNN NNNNNNNNNNNNNNNNNNNN 1
M 2101 hCoV-19/England/QEUH-A00604/2020|EPI_ISL_584893|20 ···················· ···················· ···················· ···················· NNNNNNNNNNNNNNNNNNNN NNNNNNNNNNNNNNNNNNNN 1
M 2190 hCoV-19/USA/MI-UM-10036231954/2020|EPI_ISL_590722| ···················· ···················· ···················· ···················· NNNNNNNNNNNNNNNNNNNN ···················· 1
M 2196 hCoV-19/USA/MI-UM-10036166894/2020|EPI_ISL_590729| ···················· ···················· ···················· ···················· NNNNNNNNNNNNNNNNNNNN ···················· 1
M 2201 hCoV-19/USA/MI-UM-10036188357/2020|EPI_ISL_590735| ···················· ·············A······ ···················· ···················· NNNNNNNNNNNNNNNNNNNN ···················· 1
M 2206 hCoV-19/USA/MI-UM-MH27682/2020|EPI_ISL_590752|2020 ···················· ···················· ···················· ···················· NNNNNNNNNNNNNNNNNNNN NNNNNNNNNNNNNNNNNNNN 1
M 2253 hCoV-19/Wales/PHWC-47E9C2/2020|EPI_ISL_595533|2020 ···················· ···················· ···················· ···················· NNNNNNNNNNNN········ ···················· 1
M 2256 hCoV-19/Wales/PHWC-47EA0B/2020|EPI_ISL_595537|2020 ···················· ···················· ···················· ···················· NNNNNNNNNNNN········ NNNNNNNNNNN········· 1
M 2271 hCoV-19/Wales/PHWC-47EB44/2020|EPI_ISL_595557|2020 ···················· ···················· ···················· ···················· NNNNNNNNNNNNNNNNNNNN NNNNNNNNNNNNNNNNNNNN 1
M 2290 hCoV-19/Wales/PHWC-47ED3F/2020|EPI_ISL_595583|2020 ···················· ···················· ···················· ···················· NNNNNNNNNNNNNNNNNNNN NNNNNNNNNNNNNNNNNNNN 1
M 2292 hCoV-19/Wales/PHWC-47ED5D/2020|EPI_ISL_595585|2020 ···················· ···················· ···················· ···················· NNNNNNNNNNNNNNNNNNNN ···················· 1
M 2542 hCoV-19/Australia/VIC12110/2020|EPI_ISL_591917|202 ···················· ···················· ···················· ···················· NNNNNNNNNNNNNNNNNNNN NNNNNNNNNNNNNNN····· 1
M 2545 hCoV-19/Australia/VIC12132/2020|EPI_ISL_591920|202 ···················· ···················· ···················· ···················· NNNNNNNNNNNNNNNNNNNN ···················· 1
M 2704 hCoV-19/Australia/VIC12221/2020|EPI_ISL_591933|202 ···················· ···················· ···················· ···················· NNNNNNNNNNNNNNNNNN·· NNNNNNNNNNNNNNNNNNNN 1
M 2708 hCoV-19/Australia/VIC12256/2020|EPI_ISL_591937|202 ···················· ···················· ···················· ···················· NNNNNNNNNNNNNNNNNNNN NNNNNNNNNNNNNNNNNNNN 1
M 2714 hCoV-19/Australia/VIC12274/2020|EPI_ISL_591943|202 ···················· ···················· ···················· ···················· NNNNNNNNNNNNNNNNNNNN ···················· 1
M 2727 hCoV-19/Australia/VIC12314/2020|EPI_ISL_591956|202 ···················· ···················· ···················· ···················· NNNNNNNNNNNNNNNNNNNN ···········NNNN····· 1
M 2734 hCoV-19/Australia/VIC12342/2020|EPI_ISL_591963|202 ···················· ···················· ···················· ···················· NNNNNNNNNNNNNNNNNNNN ···················· 1
M 2774 hCoV-19/Wales/PHWC-47E1D9/2020|EPI_ISL_595421|2020 ···················· ···················· ···················· ···················· NNNNNNNNNNNNNNNNNNNN NNNNNNNNN··········N 1
M 2783 hCoV-19/Wales/PHWC-47E2C7/2020|EPI_ISL_595433|2020 ···················· ···················· ···················· ···················· NNNNNNNNNNNNNNNNNNNN NNNNNNNNNNNNNNNNNNNN 1
M 2786 hCoV-19/Wales/PHWC-47E31F/2020|EPI_ISL_595438|2020 ···················· ···················· ···················· ···················· ·········NNNNNNNNNNN ···················· 1
M 2791 hCoV-19/Wales/PHWC-47E397/2020|EPI_ISL_595446|2020 ···················· ···················· ···················· ···················· NNNNNNNNNNNNNNNNNNNN NNNNNNNNNNNNNNNNNNNN 1
M 2808 hCoV-19/Wales/PHWC-47E6DA/2020|EPI_ISL_595489|2020 ···················· ···················· ···················· ···················· NNNNNNNNNNNNNNNNNNNN ···················· 1
M 2843 hCoV-19/England/SHEF-CF0B2/2020|EPI_ISL_585072|202 ···················· ···················· ···················· ···················· NNNNNNNNNNNNNNNNNNNN NNNNNNNNNNNNNNNNNNNN 1
M 2852 hCoV-19/England/SHEF-D314F/2020|EPI_ISL_585088|202 ···················· ···················· ···················· ···················· NNNNNNNNNNNNNNNNNNNN NNNNNNNNNNNNNNNNNNNN 1
M 2853 hCoV-19/England/SHEF-D318B/2020|EPI_ISL_585089|202 ···················· ···················· ···················· ···················· NNNNNNNNNNNNNNNNNNNN NNNNNNNNNNNNNNNNNNNN 1
M 2855 hCoV-19/Northern Ireland/NIRE-106F93/2020|EPI_ISL_ ···················· ···················· ···················· ···················· NNNNNNNNNNNN········ ···················· 1
M 2859 hCoV-19/Northern Ireland/NIRE-106FEE/2020|EPI_ISL_ ···················· ···················· ···················· ···················· NNNNNNNNNNNNNNNNNNNN N······NNNNNNNNNNNNN 1
M 2862 hCoV-19/Northern Ireland/NIRE-107141/2020|EPI_ISL_ ···················· ···················· ···················· ···················· NNNNNNNNNNNNNNNNNNNN NNNNNNNNNNNNNNNNNNNN 1
M 2863 hCoV-19/Northern Ireland/NIRE-10716F/2020|EPI_ISL_ ···················· ···················· ···················· ···················· NNNNNNN············· NNNN················ 1
M 2866 hCoV-19/Northern Ireland/NIRE-10719C/2020|EPI_ISL_ ···················· ···················· ···················· ···················· NNNNNNNNNNNNNNNNNNNN NNNNNNNNNNNNNNNNNNNN 1
M 2868 hCoV-19/Northern Ireland/NIRE-1071BA/2020|EPI_ISL_ ···················· ···················· ···················· ···················· NNNNNNNNNNNNNNNNNNNN NNNNNNNNNNNNNNNNNNNN 1
M 2916 hCoV-19/Australia/VIC13259/2020|EPI_ISL_592221|202 ···················· ···················· ···················· ···················· NNNNNNNNNNNNNNNNNNNN ···················· 1
M 2930 hCoV-19/Australia/VIC13428/2020|EPI_ISL_592235|202 ···················· ···················· ···················· ···················· NNNNNNNNNNNNNNNNNNNN ···················· 1
M 3017 hCoV-19/Scotland/EDB8101/2020|EPI_ISL_594828|2020- ···················· ···················· ···················· ···················· NNNNNNNNNNNNNNNNNNNN NNNNNNNNNNNNNNNNNNNN 1
M 3024 hCoV-19/Scotland/EDB8232/2020|EPI_ISL_594836|2020- ···················· ···················· ···················· ···················· NNNNNNNNNNNNNNNNNNNN NNNNNNNNNNNNNNNNNNNN 1
M 3028 hCoV-19/Scotland/EDB8291/2020|EPI_ISL_594841|2020- ···················· ···················· ···················· ···················· NNNNNNNNNNNNNNNNNNNN NNNNNNNNNNNNNNNNNNNN 1
M 3029 hCoV-19/Scotland/EDB8293/2020|EPI_ISL_594842|2020- ···················· ···················· ···················· ···················· NNNNNNNNNNNNNNNNNNNN NNNNNNNNNNNNNNNNNNNN 1
M 3031 hCoV-19/Scotland/EDB8309/2020|EPI_ISL_594847|2020- ···················· ···················· ···················· ···················· NNNNNNNNNNNNNNNNNNNN NNNNNNNNNNNNNNNNNNNN 1
M 3063 hCoV-19/Wales/PHWC-47E41C/2020|EPI_ISL_595454|2020 ···················· ···················· ···················· ···················· NNNN················ ···················· 1
M 3073 hCoV-19/Northern Ireland/NIRE-107035/2020|EPI_ISL_ ···················· ···················· ···················· ···················· NNNNNNNNNNNNNNNNNNNN NNNNNNNNNNNNNNNNNNNN 1
M 3074 hCoV-19/Northern Ireland/NIRE-107053/2020|EPI_ISL_ ···················· ···················· ···················· ···················· NNNNNNNNNNNNNNNNNNNN NNNNNNNNNNNNNNNNNNNN 1
M 3076 hCoV-19/Northern Ireland/NIRE-10709F/2020|EPI_ISL_ ···················· ···················· ···················· ···················· ···················· NNNNNNNNNNNNNNNNNNNN 1
M 3077 hCoV-19/Northern Ireland/NIRE-1070AE/2020|EPI_ISL_ ···················· ···················· ···················· ···················· NNNNNNNNNNNNNNNNNNNN NNNNNNNNNNNNNNNNNNNN 1
M 3082 hCoV-19/Northern Ireland/NIRE-1070F9/2020|EPI_ISL_ ···················· ···················· ···················· ···················· NNNNNNNNNNNNNNNNNNNN NNNNNNNNNNNNNNNNNNNN 1
M 3084 hCoV-19/Northern Ireland/NIRE-107114/2020|EPI_ISL_ ···················· ···················· ···················· ···················· ···················· NNNNNNNNNNNNNNNN···· 1
M 3088 hCoV-19/Northern Ireland/NIRE-10725D/2020|EPI_ISL_ ···················· ···················· ···················· ···················· ···················· NNNNN··············· 1
M 3089 hCoV-19/Northern Ireland/NIRE-10727B/2020|EPI_ISL_ ···················· ···················· ···················· ···················· NNNNNNNNNNNNNNNNNNNN NNNNNNNNNNNNNNNNNNNN 1
M 3092 hCoV-19/Northern Ireland/NIRE-1072E4/2020|EPI_ISL_ ···················· ···················· ···················· ···················· NNNNNNNNNNNNNNNNNNNN ···················· 1
M 3095 hCoV-19/Northern Ireland/NIRE-10732D/2020|EPI_ISL_ ···················· ···················· ···················· ···················· ···················· NNNN················ 1
M 3105 hCoV-19/Northern Ireland/NIRE-107439/2020|EPI_ISL_ ···················· ···················· ···················· ···················· NNNNNNNNNNNNNNNNNNNN ···················· 1
M 3110 hCoV-19/Northern Ireland/NIRE-107484/2020|EPI_ISL_ ···················· ···················· ···················· ···················· NNNNNNNNNNNNNNNNNNNN NNNN·······NNNNNNNNN 1
M 3112 hCoV-19/Northern Ireland/NIRE-1074A2/2020|EPI_ISL_ ···················· ···················· ···················· ···················· NNNNNNNNNNNNNNNNNNNN NNNNNNNNNNNNNNNNNNNN 1
M 3114 hCoV-19/Northern Ireland/NIRE-1074C0/2020|EPI_ISL_ ···················· ···················· ···················· ···················· ·NNNNNNNNNNNNNNNNNNN ···················· 1
M 3117 hCoV-19/Northern Ireland/NIRE-1074FD/2020|EPI_ISL_ ···················· ···················· ···················· ···················· NNNNNNNNNNNN········ ···················· 1
M 3132 hCoV-19/Northern Ireland/NIRE-10823E/2020|EPI_ISL_ ···················· ···················· ···················· ···················· NNNN················ ···················· 1
M 3133 hCoV-19/Northern Ireland/NIRE-10825C/2020|EPI_ISL_ ···················· ···················· ···················· ···················· NNNNNNNNNNNNNNNNNNNN NNNNNNNNNNNNNNNNNNNN 1
M 3134 hCoV-19/Northern Ireland/NIRE-10826B/2020|EPI_ISL_ ···················· ···················· ···················· ···················· NNNNNNNNNNNNNNNNNNNN NNNNNNNNNNNNNNNNNNNN 1
M 3297 hCoV-19/Australia/VIC15702/2020|EPI_ISL_593415|202 ···················· ···················· ···················· ···················· NNNNNNNNNNNNNNNNNNNN NNNNNNNNNNNNNNN····· 1
M 3303 hCoV-19/Australia/VIC15716/2020|EPI_ISL_593421|202 ···················· ···················· ···················· ···················· NNNNNNNNNNNNNNNNNNNN NNNNNNNNNNNNNNNNNNNN 1
M 3306 hCoV-19/Australia/VIC15724/2020|EPI_ISL_593424|202 ···················· ···················· ···················· ···················· NNNNNNNNNNNNNNNNNNNN NNNNNNNNNNNNNNNNNNNN 1
M 3307 hCoV-19/Australia/VIC15725/2020|EPI_ISL_593425|202 ···················· ······A············· ···················· ···················· ···················· NNNN················ 1
M 3309 hCoV-19/Australia/VIC15729/2020|EPI_ISL_593427|202 ···················· ···················· ···················· ···················· NNNNNNNNNNNNNNNNNNNN ···················· 1
M 3332 hCoV-19/England/SHEF-CB2ED/2020|EPI_ISL_585043|202 ···················· ···················· ···················· ···················· NNNNNNNNNNNNNNNNNNNN NNNNNNNNNNNNNNNNNNNN 1
M 3333 hCoV-19/England/SHEF-CB924/2020|EPI_ISL_585044|202 ···················· ···················· ···················· ···················· NNNNNNNNNNNNNNNNNNNN NNNNNNNNNNNNNNNNNNNN 1
M 3335 hCoV-19/England/SHEF-CC598/2020|EPI_ISL_585048|202 ···················· ···················· ···················· ···················· NNNNNNNNNNNNNNNNNNNN NNNNNNNNNNNNNNNNNNNN 1
M 3339 hCoV-19/England/SHEF-CCDAF/2020|EPI_ISL_585053|202 ···················· ·A·················· ···················· ···················· NNNNNNNNNNNNNNNNNNNN NNNNNNNNNNNNNNNNNNNN 1
M 3341 hCoV-19/England/SHEF-CCF12/2020|EPI_ISL_585055|202 ···················· ···················· ···················· ···················· NNNNNNNNNNNNNNNNNNNN NNNNNNNNNNNNNNNNNNNN 1
M 3487 hCoV-19/Australia/VIC12988/2020|EPI_ISL_592138|202 ···················· ···················· ···················· ···················· NNNNNNNNNNNNNNNNNNNN NNNNNNNNNNNNNNN····· 1
M 3498 hCoV-19/Australia/VIC15669/2020|EPI_ISL_593406|202 ···················· ···················· ···················· ···················· NNNNNNNNNNNNNNNNNNNN ···················· 1
M 3503 hCoV-19/Australia/VIC15735/2020|EPI_ISL_593431|202 ···················· ···················· ···················· ···················· NNNNNNNNNNNNNNNNNNNN NNNNNNNNNNNNNNN····· 1
M 3504 hCoV-19/Australia/VIC15738/2020|EPI_ISL_593432|202 ···················· ···················· ···················· ···················· NNNNNNNNNNNNNNNNNNNN NNNNNNNN············ 1
M 3505 hCoV-19/Australia/VIC15739/2020|EPI_ISL_593433|202 ···················· ···················· ···················· ···················· ···················· NNNN················ 1
M 3506 hCoV-19/Australia/VIC15740/2020|EPI_ISL_593434|202 ···················· ···················· ···················· ···················· NNNNNNNNNNNNNNNNNNNN NNNNNNNNN··········· 1
M 3509 hCoV-19/Australia/VIC15744/2020|EPI_ISL_593437|202 ···················· ···················· ···················· ···················· NNNNNNNNNN·········· ···················· 1
M 3513 hCoV-19/Australia/VIC15753/2020|EPI_ISL_593441|202 ···················· ···················· ···················· ···················· NNNR················ ···················· 1

<table>
<tr><td>M</td><td>3514</td><td>hCoV-19/Australia/VIC15755/2020|EPI_ISL_593442|202</td><td>····················</td><td>····················</td><td>····················</td><td>····················</td><td>·····NNNNNNNNNNNN···</td><td>NNNNN···············</td><td>1</td></tr>
<tr><td>M</td><td>3521</td><td>hCoV-19/Australia/VIC15766/2020|EPI_ISL_593449|202</td><td>····················</td><td>····················</td><td>····················</td><td>····················</td><td>NNNNNNNNNNNNNNNNNNNN</td><td>NNNNNNNNNNNNNNNNNNNN</td><td>1</td></tr>
<tr><td>M</td><td>3524</td><td>hCoV-19/Australia/VIC15776/2020|EPI_ISL_593452|202</td><td>····················</td><td>····················</td><td>····················</td><td>····················</td><td>NNNNNNNNNNNNNNNNNNNN</td><td>····················</td><td>1</td></tr>
<tr><td>M</td><td>3525</td><td>hCoV-19/Australia/VIC15778/2020|EPI_ISL_593453|202</td><td>····················</td><td>····················</td><td>····················</td><td>····················</td><td>NNNNNNNNNNNNNNNNNNNN</td><td>····················</td><td>1</td></tr>
<tr><td>M</td><td>3526</td><td>hCoV-19/Australia/VIC15779/2020|EPI_ISL_593454|202</td><td>····················</td><td>····················</td><td>····················</td><td>····················</td><td>NNNNNNNNNNNNNNNNNNNN</td><td>NNNNNNNNNNNNNNNNNNNN</td><td>1</td></tr>
<tr><td>M</td><td>3536</td><td>hCoV-19/Australia/VIC15814/2020|EPI_ISL_593464|202</td><td>····················</td><td>····················</td><td>····················</td><td>····················</td><td>NNNNNNNNNNNNNNNNNNNN</td><td>····················</td><td>1</td></tr>
<tr><td>M</td><td>3545</td><td>hCoV-19/Australia/VIC15837/2020|EPI_ISL_593473|202</td><td>····················</td><td>····················</td><td>····················</td><td>····················</td><td>NNNNNNNNNNNNNNNNNNNN</td><td>····················</td><td>1</td></tr>
<tr><td>M</td><td>3548</td><td>hCoV-19/Scotland/CVR4936/2020|EPI_ISL_594702|2020-</td><td>····················</td><td>····················</td><td>····················</td><td>····················</td><td>····················</td><td>NNNN··········NN····</td><td>1</td></tr>
<tr><td>M</td><td>3558</td><td>hCoV-19/Scotland/CVR4966/2020|EPI_ISL_594730|2020-</td><td>····················</td><td>····················</td><td>····················</td><td>····················</td><td>···········N··NN····</td><td>NN·······NN·········</td><td>1</td></tr>
<tr><td>M</td><td>3561</td><td>hCoV-19/Scotland/CVR4974/2020|EPI_ISL_594737|2020-</td><td>····················</td><td>····················</td><td>····················</td><td>····················</td><td>··············NN····</td><td>NNNN················</td><td>1</td></tr>
<tr><td>M</td><td>3601</td><td>hCoV-19/England/SHEF-C3CD8/2020|EPI_ISL_585019|202</td><td>····················</td><td>····················</td><td>····················</td><td>····················</td><td>NNNNNNNNNNNNNNNNNNNN</td><td>NNNNNNNNNNNNNNNNNNNN</td><td>1</td></tr>
<tr><td>M</td><td>3605</td><td>hCoV-19/England/SHEF-C6385/2020|EPI_ISL_585025|202</td><td>····················</td><td>····················</td><td>····················</td><td>····················</td><td>NNNNNNNNNNNNNNNNNNNN</td><td>NNNNNNNNNNNNNNNNNNNN</td><td>1</td></tr>
<tr><td>M</td><td>3609</td><td>hCoV-19/England/SHEF-C79BF/2020|EPI_ISL_585031|202</td><td>····················</td><td>····················</td><td>····················</td><td>····················</td><td>NNNNNNNNNNNNNNNNNNNN</td><td>NNNNNNNNNNNNNNNNNNNN</td><td>1</td></tr>
<tr><td>M</td><td>3610</td><td>hCoV-19/England/SHEF-C79DD/2020|EPI_ISL_585032|202</td><td>····················</td><td>····················</td><td>····················</td><td>····················</td><td>NNNNNNNNNNNNNNNNNNNN</td><td>NNNNNNNNNNNNNNNNNNNN</td><td>1</td></tr>
<tr><td>M</td><td>3792</td><td>hCoV-19/Australia/VIC15839/2020|EPI_ISL_593474|202</td><td>····················</td><td>····················</td><td>····················</td><td>····················</td><td>NNNNNNNNNNNNNNNNNNNN</td><td>····················</td><td>1</td></tr>
<tr><td>M</td><td>3794</td><td>hCoV-19/Australia/VIC3512/2020|EPI_ISL_593476|2020</td><td>····················</td><td>····················</td><td>····················</td><td>····················</td><td>NNNN················</td><td>····················</td><td>1</td></tr>
<tr><td>M</td><td>3830</td><td>hCoV-19/Scotland/QEUH-A4906B/2020|EPI_ISL_595951|2</td><td>····················</td><td>····················</td><td>····················</td><td>····················</td><td>NNNNNNNNNNNNNNNNNNNN</td><td>NNNNN···············</td><td>1</td></tr>
<tr><td>M</td><td>3840</td><td>hCoV-19/Scotland/QEUH-A4AAC6/2020|EPI_ISL_595970|2</td><td>····················</td><td>····················</td><td>····················</td><td>····················</td><td>NNNNNNNNNNNNNNNNNNNN</td><td>NNNNNNNNNNNNNNNNNNNN</td><td>1</td></tr>
<tr><td>M</td><td>3861</td><td>hCoV-19/Wales/PHWC-47D4D1/2020|EPI_ISL_586152|2020</td><td>····················</td><td>····················</td><td>····················</td><td>····················</td><td>NNNNNNNNNNNNNNNNNNNN</td><td>NNNNNNNNNNNNNNNNNNNN</td><td>1</td></tr>
<tr><td>M</td><td>3862</td><td>hCoV-19/Wales/PHWC-47D4E0/2020|EPI_ISL_586153|2020</td><td>····················</td><td>····················</td><td>····················</td><td>····················</td><td>NNNNNNNNNNNNNNNNNN··</td><td>NNNNNNNNNNNNNNN··NNN</td><td>1</td></tr>
<tr><td>M</td><td>3864</td><td>hCoV-19/Wales/PHWC-47D547/2020|EPI_ISL_586158|2020</td><td>····················</td><td>····················</td><td>····················</td><td>····················</td><td>NNNNNNNNNNNNNNNNNNNN</td><td>····················</td><td>1</td></tr>
<tr><td>M</td><td>3867</td><td>hCoV-19/Wales/PHWC-47D5ED/2020|EPI_ISL_586167|2020</td><td>····················</td><td>····················</td><td>····················</td><td>····················</td><td>NNNNNNNNNNNNNNNNNNNN</td><td>··············NNNNNN</td><td>1</td></tr>
<tr><td>M</td><td>3868</td><td>hCoV-19/Wales/PHWC-47D5FC/2020|EPI_ISL_586168|2020</td><td>····················</td><td>····················</td><td>····················</td><td>····················</td><td>NNNNNNNNNNNNNNNNNNNN</td><td>N···················</td><td>1</td></tr>
<tr><td>M</td><td>3871</td><td>hCoV-19/Wales/PHWC-47D653/2020|EPI_ISL_586172|2020</td><td>····················</td><td>····················</td><td>····················</td><td>····················</td><td>NNNNNNNNNNNNNNNNNNNN</td><td>NNNNNNNNNNNNNNNNNNNN</td><td>1</td></tr>
<tr><td>M</td><td>3875</td><td>hCoV-19/Wales/PHWC-47D6AE/2020|EPI_ISL_586177|2020</td><td>····················</td><td>····················</td><td>····················</td><td>····················</td><td>NNNNNNNNNNNNNNNNNNNN</td><td>NNNNNNNNNNNNNNNNNNNN</td><td>1</td></tr>
<tr><td>M</td><td>3876</td><td>hCoV-19/Wales/PHWC-47D6BD/2020|EPI_ISL_586178|2020</td><td>····················</td><td>····················</td><td>····················</td><td>····················</td><td>NNNNNNNNNNNNNNNNNNNN</td><td>NNNNNNNNNNNNNNNNNNNN</td><td>1</td></tr>
<tr><td>M</td><td>3878</td><td>hCoV-19/Wales/PHWC-47D6DB/2020|EPI_ISL_586180|2020</td><td>····················</td><td>····················</td><td>····················</td><td>····················</td><td>NNNNNNNNNNNNNNNNNNNN</td><td>····················</td><td>1</td></tr>
<tr><td>M</td><td>3886</td><td>hCoV-19/Wales/PHWC-47D79C/2020|EPI_ISL_586190|2020</td><td>····················</td><td>····················</td><td>····················</td><td>····················</td><td>NNNNNNNNNNNNNNNNNNNN</td><td>····················</td><td>1</td></tr>
<tr><td>M</td><td>3887</td><td>hCoV-19/Wales/PHWC-47D7AB/2020|EPI_ISL_586191|2020</td><td>····················</td><td>····················</td><td>····················</td><td>····················</td><td>NNNNNNNNNNNNNNNNNNNN</td><td>NNNNNNNNNNNNNNNNNNNN</td><td>1</td></tr>
<tr><td>M</td><td>3891</td><td>hCoV-19/Wales/PHWC-47D820/2020|EPI_ISL_586198|2020</td><td>····················</td><td>····················</td><td>····················</td><td>····················</td><td>NNNNNNNNNNNNNNNNNNNN</td><td>NNNNNNNNNNNNNNNNNNNN</td><td>1</td></tr>
<tr><td>M</td><td>3892</td><td>hCoV-19/Wales/PHWC-47D84E/2020|EPI_ISL_586199|2020</td><td>····················</td><td>····················</td><td>····················</td><td>····················</td><td>NNNNNNNNNNNNNNNNNNNN</td><td>NNNNNNNNNNNNNNNNNNNN</td><td>1</td></tr>
<tr><td>M</td><td>3893</td><td>hCoV-19/Wales/PHWC-47D8B7/2020|EPI_ISL_586204|2020</td><td>····················</td><td>····················</td><td>····················</td><td>····················</td><td>NNNNNNNNNNNNNNNNNNNN</td><td>····················</td><td>1</td></tr>
<tr><td>M</td><td>3898</td><td>hCoV-19/Wales/PHWC-47D93C/2020|EPI_ISL_586211|2020</td><td>····················</td><td>····················</td><td>····················</td><td>····················</td><td>NNNNNNNNNNNNNNNNNNNN</td><td>NNNNNNNNNNNNNNNNNNNN</td><td>1</td></tr>
<tr><td>M</td><td>3900</td><td>hCoV-19/Wales/PHWC-47D95A/2020|EPI_ISL_586213|2020</td><td>····················</td><td>·A··················</td><td>····················</td><td>····················</td><td>NNNNNNNNNNNNNNNNNNNN</td><td>····················</td><td>1</td></tr>
<tr><td>M</td><td>3951</td><td>hCoV-19/Australia/VIC12525/2020|EPI_ISL_592015|202</td><td>····················</td><td>····················</td><td>····················</td><td>····················</td><td>····················</td><td>NNNNNNNNNNNNNNNNNNNN</td><td>1</td></tr>
<tr><td>M</td><td>3956</td><td>hCoV-19/Australia/VIC12536/2020|EPI_ISL_592020|202</td><td>····················</td><td>····················</td><td>····················</td><td>····················</td><td>NNNNNNNNNNNNNNNNNNNN</td><td>NNNNNNNNNNNNNNNNNNNN</td><td>1</td></tr>
<tr><td>M</td><td>3958</td><td>hCoV-19/Australia/VIC15441/2020|EPI_ISL_593307|202</td><td>····················</td><td>····················</td><td>····················</td><td>····················</td><td>NNNNNNNNNNNNNNNNNNNN</td><td>NNNNNNNNNNNNNNNNNNNN</td><td>1</td></tr>
<tr><td>M</td><td>3991</td><td>hCoV-19/Australia/VIC15510/2020|EPI_ISL_593342|202</td><td>····················</td><td>····················</td><td>····················</td><td>····················</td><td>NNNNNNNNNNNNNNNNNNNN</td><td>····················</td><td>1</td></tr>
<tr><td>M</td><td>4002</td><td>hCoV-19/England/BIRM-65A29/2020|EPI_ISL_594602|202</td><td>····················</td><td>····················</td><td>····················</td><td>····················</td><td>NNNNNNNNNNNNNNNNNNNN</td><td>NNNNNNNNNNNNNNNNNNNN</td><td>1</td></tr>
<tr><td>M</td><td>4004</td><td>hCoV-19/England/BIRM-65A47/2020|EPI_ISL_594604|202</td><td>····················</td><td>····················</td><td>····················</td><td>····················</td><td>NNNNNNNNNNNNNNNNNNNN</td><td>NNNNNNNNNNNNNNNNNNNN</td><td>1</td></tr>
<tr><td>M</td><td>4005</td><td>hCoV-19/England/BIRM-65A56/2020|EPI_ISL_594605|202</td><td>····················</td><td>····················</td><td>····················</td><td>····················</td><td>NNNNNNNNNNNNNNNNNNNN</td><td>NNNNNNNNNNNNNNNNNNNN</td><td>1</td></tr>
<tr><td>M</td><td>4010</td><td>hCoV-19/England/BIRM-65F57/2020|EPI_ISL_594610|202</td><td>····················</td><td>····················</td><td>····················</td><td>····················</td><td>NNNNNNNNNNNNNNNNNNNN</td><td>NNNNNNNNNNNNNNNNNNNN</td><td>1</td></tr>
<tr><td>M</td><td>4011</td><td>hCoV-19/England/BIRM-65F84/2020|EPI_ISL_594611|202</td><td>····················</td><td>····················</td><td>····················</td><td>····················</td><td>NNNNNNNNNNNNNNNNNNNN</td><td>NNNNNNNNNNNNNNNNNNNN</td><td>1</td></tr>
<tr><td>M</td><td>4012</td><td>hCoV-19/England/BIRM-65FB1/2020|EPI_ISL_594612|202</td><td>····················</td><td>····················</td><td>····················</td><td>····················</td><td>NNNNNNNNNNNNNNNNNNNN</td><td>NNNNNNNNNNNNNNNNNNNN</td><td>1</td></tr>
<tr><td>M</td><td>4013</td><td>hCoV-19/England/BIRM-66053/2020|EPI_ISL_594614|202</td><td>····················</td><td>····················</td><td>····················</td><td>····················</td><td>NNNNNNNNNNNNNNNNNNNN</td><td>NNNNNNNNNNNNNNNNNNNN</td><td>1</td></tr>
<tr><td>M</td><td>4014</td><td>hCoV-19/England/BIRM-66071/2020|EPI_ISL_594615|202</td><td>····················</td><td>····················</td><td>····················</td><td>····················</td><td>NNNNNNNNNNNNNNNNNNNN</td><td>NNNNNNNNNNNNNNNNNNNN</td><td>1</td></tr>
<tr><td>M</td><td>4015</td><td>hCoV-19/England/BIRM-66080/2020|EPI_ISL_594616|202</td><td>····················</td><td>····················</td><td>····················</td><td>····················</td><td>NNNNNNNNNNNNNNNNNNNN</td><td>NNNNNNNNNNNNNNNNNNNN</td><td>1</td></tr>
<tr><td>M</td><td>4026</td><td>hCoV-19/Scotland/CVR4851/2020|EPI_ISL_594651|2020-</td><td>····················</td><td>····················</td><td>····················</td><td>····················</td><td>····················</td><td>NNNN···NN···········</td><td>1</td></tr>
<tr><td>M</td><td>4041</td><td>hCoV-19/Scotland/CVR4917/2020|EPI_ISL_594683|2020-</td><td>····················</td><td>····················</td><td>····················</td><td>····················</td><td>····················</td><td>NN·N···NN····N······</td><td>1</td></tr>
<tr><td>M</td><td>4042</td><td>hCoV-19/Scotland/QEUH-A3153A/2020|EPI_ISL_595900|2</td><td>····················</td><td>····················</td><td>····················</td><td>····················</td><td>NNNNNNNNNNNNNNNNNNNN</td><td>NNNNN···············</td><td>1</td></tr>
<tr><td>M</td><td>4043</td><td>hCoV-19/England/QEUH-A31576/2020|EPI_ISL_595902|20</td><td>····················</td><td>····················</td><td>····················</td><td>····················</td><td>NNNNN····NNN········</td><td>····················</td><td>1</td></tr>
<tr><td>M</td><td>4048</td><td>hCoV-19/England/QEUH-A31770/2020|EPI_ISL_595908|20</td><td>····················</td><td>····················</td><td>····················</td><td>····················</td><td>NNNNNNNNNNNNNNNNNNNN</td><td>····················</td><td>1</td></tr>
<tr><td>M</td><td>4053</td><td>hCoV-19/England/QEUH-A317DA/2020|EPI_ISL_595913|20</td><td>····················</td><td>····················</td><td>····················</td><td>····················</td><td>NNNNNNNNNNNNNNNNNNNN</td><td>NNNNNNNNNNNNNNNNNNNN</td><td>1</td></tr>
<tr><td>M</td><td>4056</td><td>hCoV-19/England/QEUH-A48CC2/2020|EPI_ISL_595917|20</td><td>····················</td><td>····················</td><td>····················</td><td>····················</td><td>NNNNNNNNNNNNNNNN····</td><td>NNNNN···············</td><td>1</td></tr>
<tr><td>M</td><td>4057</td><td>hCoV-19/England/QEUH-A48CD1/2020|EPI_ISL_595918|20</td><td>····················</td><td>····················</td><td>····················</td><td>····················</td><td>NNNNNNN·············</td><td>NNNN················</td><td>1</td></tr>
<tr><td>M</td><td>4058</td><td>hCoV-19/England/QEUH-A48CFF/2020|EPI_ISL_595920|20</td><td>····················</td><td>····················</td><td>····················</td><td>····················</td><td>NNNNNNNNNNNNNNNNNNNN</td><td>NNNNNNNNNNNNNNNN·N·N</td><td>1</td></tr>
<tr><td>M</td><td>4059</td><td>hCoV-19/Scotland/QEUH-A48D29/2020|EPI_ISL_595922|2</td><td>····················</td><td>····················</td><td>····················</td><td>····················</td><td>NNNNNNNNNNNNNNNNNNNN</td><td>NNNNNNNNNNNNNNNNNNNN</td><td>1</td></tr>
<tr><td>M</td><td>4069</td><td>hCoV-19/Scotland/QEUH-A48F6F/2020|EPI_ISL_595940|2</td><td>····················</td><td>····················</td><td>····················</td><td>····················</td><td>NNNNNNNNNNNNN·······</td><td>····················</td><td>1</td></tr>
<tr><td>M</td><td>4082</td><td>hCoV-19/Scotland/QEUH-A4C587/2020|EPI_ISL_595979|2</td><td>····················</td><td>····················</td><td>····················</td><td>····················</td><td>NNNNNNNNNNNNNNNNNNNN</td><td>NNNNNNNNNNNNNNNNNNNN</td><td>1</td></tr>
<tr><td>M</td><td>4083</td><td>hCoV-19/Scotland/QEUH-A4C596/2020|EPI_ISL_595980|2</td><td>····················</td><td>····················</td><td>····················</td><td>····················</td><td>NNN·················</td><td>NNNNNNNNNNNNNNNN····</td><td>1</td></tr>
<tr><td>M</td><td>4084</td><td>hCoV-19/England/QEUH-A4C5A5/2020|EPI_ISL_595981|20</td><td>····················</td><td>····················</td><td>····················</td><td>····················</td><td>NNNNNNNNNNNNNNNNNNNN</td><td>NNNNN···············</td><td>1</td></tr>
<tr><td>M</td><td>4086</td><td>hCoV-19/Scotland/QEUH-A4C5D2/2020|EPI_ISL_595983|2</td><td>····················</td><td>····················</td><td>····················</td><td>····················</td><td>NNNNNNNNNNNNNNNNNNNN</td><td>NNNNNNNNNNNNNNNNNNNN</td><td>1</td></tr>
<tr><td>M</td><td>4087</td><td>hCoV-19/England/QEUH-A4C684/2020|EPI_ISL_595987|20</td><td>····················</td><td>·A··················</td><td>····················</td><td>····················</td><td>N···················</td><td>····················</td><td>1</td></tr>
<tr><td>M</td><td>4091</td><td>hCoV-19/Scotland/QEUH-A4C6FD/2020|EPI_ISL_595992|2</td><td>····················</td><td>····················</td><td>····················</td><td>····················</td><td>NNNNNNNNNNNNNNNNNNNN</td><td>NNNNNNNNNNN·········</td><td>1</td></tr>
<tr><td>M</td><td>4096</td><td>hCoV-19/England/QEUH-A4C763/2020|EPI_ISL_595999|20</td><td>····················</td><td>····················</td><td>····················</td><td>····················</td><td>NNNNNNNNNNNNNNNNNNNN</td><td>NNNNNNNNNNNNNNNNNNNN</td><td>1</td></tr>
<tr><td>M</td><td>4098</td><td>hCoV-19/Wales/PHWC-47D88A/2020|EPI_ISL_586202|2020</td><td>····················</td><td>····················</td><td>····················</td><td>····················</td><td>NNNNNNNNNNNNNNNNNNNN</td><td>NNNNNNNNNNNNNNNNNNNN</td><td>1</td></tr>
<tr><td>M</td><td>4099</td><td>hCoV-19/Wales/PHWC-47D969/2020|EPI_ISL_586214|2020</td><td>····················</td><td>····················</td><td>····················</td><td>····················</td><td>NNNNNNNNNNNNNNNNNNN·</td><td>····················</td><td>1</td></tr>
<tr><td>M</td><td>4104</td><td>hCoV-19/Wales/PHWC-47D9D2/2020|EPI_ISL_586221|2020</td><td>····················</td><td>····················</td><td>····················</td><td>····················</td><td>NNNNNNNNNNNNNNNNNNNN</td><td>····················</td><td>1</td></tr>
<tr><td>M</td><td>4107</td><td>hCoV-19/Wales/PHWC-47DA0C/2020|EPI_ISL_586224|2020</td><td>····················</td><td>····················</td><td>····················</td><td>····················</td><td>NNNNNNNNNNNNNNNNNNNN</td><td>····················</td><td>1</td></tr>
<tr><td>M</td><td>4110</td><td>hCoV-19/Wales/PHWC-47DA75/2020|EPI_ISL_586231|2020</td><td>····················</td><td>····················</td><td>····················</td><td>····················</td><td>····················</td><td>NNNNNNNNNNNNNNN·····</td><td>1</td></tr>
<tr><td>M</td><td>4112</td><td>hCoV-19/Wales/PHWC-47DAA2/2020|EPI_ISL_586234|2020</td><td>····················</td><td>····················</td><td>····················</td><td>····················</td><td>NNNNNNNNNNNNNNNNNNNN</td><td>····················</td><td>1</td></tr>
<tr><td>M</td><td>4114</td><td>hCoV-19/Wales/PHWC-47DADF/2020|EPI_ISL_586237|2020</td><td>····················</td><td>····················</td><td>····················</td><td>····················</td><td>NNNNNNNNNNNNNNNNNNNN</td><td>····················</td><td>1</td></tr>
<tr><td>M</td><td>4115</td><td>hCoV-19/Wales/PHWC-47DAEE/2020|EPI_ISL_586238|2020</td><td>····················</td><td>····················</td><td>····················</td><td>····················</td><td>NNNNNNNNNNNNNNNNNNNN</td><td>·················NNN</td><td>1</td></tr>
<tr><td>M</td><td>4268</td><td>hCoV-19/Australia/VIC12602/2020|EPI_ISL_592034|202</td><td>····················</td><td>····················</td><td>····················</td><td>····················</td><td>NNNNNNNNNNNNNNNNNNNN</td><td>····················</td><td>1</td></tr>
<tr><td>M</td><td>4278</td><td>hCoV-19/Australia/VIC12650/2020|EPI_ISL_592044|202</td><td>····················</td><td>····················</td><td>····················</td><td>····················</td><td>NNNNNNNNNNNNNNNNNNNN</td><td>NNNNNNNNNNN·········</td><td>1</td></tr>
<tr><td>M</td><td>4285</td><td>hCoV-19/Australia/VIC12671/2020|EPI_ISL_592051|202</td><td>····················</td><td>····················</td><td>····················</td><td>····················</td><td>NNNNNNNNNNNNNNN·····</td><td>····················</td><td>1</td></tr>
<tr><td>M</td><td>4300</td><td>hCoV-19/Australia/VIC12848/2020|EPI_ISL_592090|202</td><td>····················</td><td>····················</td><td>····················</td><td>····················</td><td>NNNNNNNNNNNNNNNNNNNN</td><td>····················</td><td>1</td></tr>
<tr><td>M</td><td>4307</td><td>hCoV-19/Australia/VIC15551/2020|EPI_ISL_593358|202</td><td>····················</td><td>····················</td><td>····················</td><td>····················</td><td>NNNNNNNNNNNNNNNNNNNN</td><td>NNNNNNNNNNNNNNNNNNNN</td><td>1</td></tr>
<tr><td>M</td><td>4318</td><td>hCoV-19/Australia/VIC15580/2020|EPI_ISL_593369|202</td><td>····················</td><td>····················</td><td>····················</td><td>····················</td><td>NNNNNNNNNNNNNNNNNNNN</td><td>····················</td><td>1</td></tr>
<tr><td>M</td><td>4319</td><td>hCoV-19/Australia/VIC15593/2020|EPI_ISL_593370|202</td><td>····················</td><td>····················</td><td>····················</td><td>····················</td><td>NNNNNNNNNNNNNNNNNNNN</td><td>····················</td><td>1</td></tr>
</table>

| | | | | | | | | |
|---|---|---|---|---|---|---|---|---|
| M | 4325 hCoV-19/Australia/VIC15614/2020\|EPI_ISL_593376\|202 | ···················· | ···················· | ···················· | ···················· | ···················· | NNNN················ | 1 |
| M | 4329 hCoV-19/Australia/VIC15620/2020\|EPI_ISL_593380\|202 | ···················· | ···················· | ···················· | ···················· | ·············NNNNNNN | ···················· | 1 |
| M | 4330 hCoV-19/Australia/VIC15621/2020\|EPI_ISL_593381\|202 | ···················· | ···················· | ···················· | ···················· | NNNN················ | NNNNNNNNN··········· | 1 |
| M | 4331 hCoV-19/Australia/VIC15622/2020\|EPI_ISL_593382\|202 | ···················· | ···················· | ···················· | ···················· | NNNNNNNNNNNN········ | ···················· | 1 |
| M | 4338 hCoV-19/Australia/VIC15634/2020\|EPI_ISL_593389\|202 | ···················· | ···················· | ···················· | ···················· | NNNNNNNNNNNNNNNNNNN· | ···················· | 1 |
| M | 4339 hCoV-19/Australia/VIC15635/2020\|EPI_ISL_593390\|202 | ···················· | ···················· | ···················· | ···················· | NNNNNNNNNNNNNNNNNNNN | ···················· | 1 |
| M | 4342 hCoV-19/Australia/VIC15642/2020\|EPI_ISL_593393\|202 | ···················· | ···················· | ···················· | ···················· | NNNNNNNNNNNNNNNNNNNN | NNNNNNNNNNNNNNNNNNNN | 1 |
| M | 4344 hCoV-19/Australia/VIC15647/2020\|EPI_ISL_593395\|202 | ···················· | ···················· | ···················· | ···················· | NNNNNNNNNNN········· | ···················· | 1 |
| M | 4345 hCoV-19/Australia/VIC15648/2020\|EPI_ISL_593396\|202 | ···················· | ···················· | ···················· | ···················· | NNNNNNNNNNNNNNNNNNNN | ···················· | 1 |
| M | 4466 hCoV-19/Australia/VIC12732/2020\|EPI_ISL_592067\|202 | ···················· | ···················· | ···················· | ···················· | NNNNNNNNNNNNNNNNNNNN | NNNNNNNNNNNNNNNNNNNN | 1 |
| M | 4468 hCoV-19/Australia/VIC12738/2020\|EPI_ISL_592069\|202 | ···················· | ···················· | ···················· | ···················· | NNNN················ | NNNNNNNNNNNNNNNNNNNN | 1 |
| M | 4471 hCoV-19/Australia/VIC12759/2020\|EPI_ISL_592072\|202 | ···················· | ···················· | ···················· | ···················· | NNNN················ | ···················· | 1 |
| M | 4477 hCoV-19/Australia/VIC12780/2020\|EPI_ISL_592078\|202 | ···················· | ···················· | ···················· | ···················· | ·NNNNNNNNNNNNNNNNNNN | ···················· | 1 |
| M | 4480 hCoV-19/Australia/VIC12797/2020\|EPI_ISL_592081\|202 | ···················· | ···················· | ···················· | ···················· | NNNNNNNNNNNNNNNNNNNN | NNN·NNNNNNNNNNN····· | 1 |
| M | 4496 hCoV-19/Australia/VIC12876/2020\|EPI_ISL_592099\|202 | ···················· | ···················· | ···················· | ···················· | NNNNNNNNNNNNNNNNNNNN | NNNNNNNNNNNNNNN····· | 1 |
| M | 4506 hCoV-19/Australia/VIC15240/2020\|EPI_ISL_593213\|202 | ···················· | ···················· | ···················· | ···················· | NNNNNNNNNNNNNNNNNNNN | ···················· | 1 |
| M | 4509 hCoV-19/Australia/VIC15244/2020\|EPI_ISL_593216\|202 | ···················· | ···················· | ···················· | ···················· | NNNNNNNNNNNNNNNNNNNN | NNNNNNNNNNNNNNNNNNNN | 1 |
| M | 4513 hCoV-19/Australia/VIC15251/2020\|EPI_ISL_593220\|202 | ···················· | ···················· | ···················· | ···················· | NNNNNNNNNNNNNNNNNNNN | ···················· | 1 |
| M | 4526 hCoV-19/Australia/VIC15655/2020\|EPI_ISL_593398\|202 | ···················· | ···················· | ···················· | ···················· | NNNNNNNNNNNNNNNNNNNN | ···················· | 1 |
| M | 4527 hCoV-19/Australia/VIC15658/2020\|EPI_ISL_593399\|202 | ···················· | ···················· | ···················· | ···················· | NNNNNNNNNNNNNNNNNNNN | ··············N····· | 1 |
| M | 4528 hCoV-19/England/BIRM-64635/2020\|EPI_ISL_594500\|202 | ···················· | ·A·················· | ···················· | ···················· | NNNNNNNNNNNNNNNNNNNN | NNNNNNNNNNNNNNNNNNNN | 1 |
| M | 4535 hCoV-19/England/BIRM-6487B/2020\|EPI_ISL_594519\|202 | ···················· | ···················· | ···················· | ···················· | NNNNNNNNNNNNNNNNNNNN | NNNNNNNNNNNNNNNNNNNN | 1 |
| M | 4536 hCoV-19/England/BIRM-648A8/2020\|EPI_ISL_594521\|202 | ···················· | ···················· | ···················· | ···················· | NNNNNNNNNNNNNNNNNNNN | NNNNNNNNNNNNNNNNNNNN | 1 |
| M | 4538 hCoV-19/England/BIRM-64E6A/2020\|EPI_ISL_594524\|202 | ···················· | ·A·················· | ···················· | ···················· | NNNNNNNNNNNNNNNNNNNN | NNNNNNNNNNNNNNNNNNNN | 1 |
| M | 4546 hCoV-19/England/BIRM-64F58/2020\|EPI_ISL_594535\|202 | ···················· | ···················· | ···················· | ···················· | NNNNNNNNNNNNNNNNNNNN | NNNNNNNNNNNNNNNNNNNN | 1 |
| M | 4547 hCoV-19/England/BIRM-64F94/2020\|EPI_ISL_594536\|202 | ···················· | ···················· | ···················· | ···················· | NNNNNNNNNNNNNNNNNNNN | NNNNNNNNNNNNNNNNNNNN | 1 |
| M | 4548 hCoV-19/England/BIRM-64FA3/2020\|EPI_ISL_594537\|202 | ···················· | ···················· | ···················· | ···················· | NNNNNNNNNNNNNNNNNNNN | NNNNNNNNNNNNNNNNNNNN | 1 |
| M | 4552 hCoV-19/England/BIRM-652D6/2020\|EPI_ISL_594545\|202 | ···················· | ···················· | ···················· | ···················· | NNNNNNNNNNNNNNNNNNNN | NNNNNNNNNNNNNNNNNNNN | 1 |
| M | 4561 hCoV-19/England/BIRM-65494/2020\|EPI_ISL_594559\|202 | ···················· | ···················· | ···················· | ···················· | NNNNNNNNNNNNNNNNNNNN | NNNNNNNNNNNNNNNNNNNN | 1 |
| M | 4562 hCoV-19/England/BIRM-654A3/2020\|EPI_ISL_594560\|202 | ···················· | ·A·················· | ···················· | ···················· | NNNNNNNNNNNNNNNNNNNN | NNNNNNNNNNNNNNNNNNNN | 1 |
| M | 4579 hCoV-19/England/QEUH-A02972/2020\|EPI_ISL_595844\|20 | ···················· | ···················· | ···················· | ···················· | NNNNNNNNNNNNNNNNNNNN | NNNNNNNNNNNNNNNNNNNN | 1 |
| M | 4580 hCoV-19/England/QEUH-A02981/2020\|EPI_ISL_595845\|20 | ···················· | ···················· | ···················· | ···················· | NNNNNNNNNNNNNNNNNNNN | NNNNNNNNNNNNNNNNNNNN | 1 |
| M | 4581 hCoV-19/England/QEUH-A30F4C/2020\|EPI_ISL_595848\|20 | ···················· | ···················· | ···················· | ···················· | NNNNNNNNNNNNNNNNNNNN | NNNNNNNNNNNNNNNNNNNN | 1 |
| M | 4584 hCoV-19/England/QEUH-A31039/2020\|EPI_ISL_595855\|20 | ···················· | ···················· | ···················· | ···················· | NNNNNNNNNNNNNNNNNNNN | ···················· | 1 |
| M | 4586 hCoV-19/England/QEUH-A31075/2020\|EPI_ISL_595858\|20 | ···················· | ···················· | ···················· | ···················· | NNNNNNNNNNNNNNNNNNNN | NNNNN··············· | 1 |
| M | 4590 hCoV-19/England/QEUH-A31109/2020\|EPI_ISL_595866\|20 | ···················· | ···················· | ···················· | ···················· | NNNNNNNNNNNNNNNNNNNN | NNNNNNNNNNNNNNNNNNNN | 1 |
| M | 4595 hCoV-19/England/QEUH-A3128E/2020\|EPI_ISL_595876\|20 | ···················· | ···················· | ···················· | ···················· | NNNNNNNNNNNNNNNNNNNN | NNNNNNNNNNNNNNNNNNNN | 1 |
| M | 4642 hCoV-19/Wales/PHWC-16F8AC/2020\|EPI_ISL_586032\|2020 | ···················· | ···················· | ···················· | ···················· | NNNNNNNNNNNNNNNNNNNN | NNNNNNNNNNNNNNNNNNNN | 1 |
| M | 4643 hCoV-19/Wales/PHWC-16F8BB/2020\|EPI_ISL_586033\|2020 | ···················· | ···················· | ···················· | ···················· | NNNNNNNNNNNNNNNNNNNN | ····NNNNNNNNNNNNNNNN | 1 |
| M | 4644 hCoV-19/Wales/PHWC-16F8CA/2020\|EPI_ISL_586034\|2020 | ···················· | ···················· | ···················· | ···················· | NNNNNNNNNNNNNNNNNNNN | NNNNNNNNNNNNNNNNNNNN | 1 |
| M | 4645 hCoV-19/Wales/PHWC-16F8F7/2020\|EPI_ISL_586037\|2020 | ···················· | ···················· | ···················· | ···················· | NNNNNNNNNNNNNNNNNNNN | NNNNNNNNNNNNNNNNNNNN | 1 |
| M | 4648 hCoV-19/Wales/PHWC-47CD22/2020\|EPI_ISL_586040\|2020 | ···················· | ···················· | ···················· | ···················· | NNNNNNNNNNNNNNNNNNNN | NNNNNNNNNNNNNNNNNNNN | 1 |
| M | 4653 hCoV-19/Wales/PHWC-47CD9B/2020\|EPI_ISL_586047\|2020 | ···················· | ···················· | ···················· | ···················· | ···················· | NNNNNNNNNNNNNNNN···· | 1 |
| M | 4657 hCoV-19/Wales/PHWC-47CDF5/2020\|EPI_ISL_586052\|2020 | ···················· | ···················· | ···················· | ···················· | NNNNNNNNNNNNNNNNNNNN | NNNNNNNNNNNNNNNNNNNN | 1 |
| M | 4658 hCoV-19/Wales/PHWC-47CE10/2020\|EPI_ISL_586054\|2020 | ···················· | ···················· | ···················· | ···················· | NNNNNNNNNNNNNNNNNNNN | ···················· | 1 |
| M | 4659 hCoV-19/Wales/PHWC-47CE3E/2020\|EPI_ISL_586056\|2020 | ···················· | ···················· | ···················· | ···················· | NNNNNNNNNNNNNNNNNNNN | ···················· | 1 |
| M | 4663 hCoV-19/Wales/PHWC-47CEA7/2020\|EPI_ISL_586063\|2020 | ···················· | ···················· | ···················· | ···················· | ···················· | NNNNNNNNNNNNNNNNN··· | 1 |
| M | 4672 hCoV-19/Wales/PHWC-47CF59/2020\|EPI_ISL_586074\|2020 | ···················· | ···················· | ···················· | ···················· | NNNNNNNNNNNNNNNNNNNN | ···················· | 1 |
| M | 4678 hCoV-19/Wales/PHWC-47CFD1/2020\|EPI_ISL_586082\|2020 | ···················· | ···················· | ···················· | ···················· | NNNNNNNNNNNNNNNNNNNN | NNNNNNNNN··········· | 1 |
| M | 4683 hCoV-19/Wales/PHWC-47D073/2020\|EPI_ISL_586092\|2020 | ···················· | ···················· | ···················· | ···················· | NNNNNNNNNNNNNNNNNNNN | NNNNNNNNNNNNNNNNNNNN | 1 |
| M | 4685 hCoV-19/Wales/PHWC-47D0A0/2020\|EPI_ISL_586094\|2020 | ···················· | ···················· | ···················· | ···················· | NNNNNNNNNNNNNNNNNNNN | ···················· | 1 |
| M | 4686 hCoV-19/Wales/PHWC-47D0CE/2020\|EPI_ISL_586096\|2020 | ···················· | ···················· | ···················· | ···················· | NNNNNNNNNNNNNNNNNNNN | NNNNNNNNNNNNNNNNNNNN | 1 |
| M | 4687 hCoV-19/Wales/PHWC-47D0DD/2020\|EPI_ISL_586097\|2020 | ···················· | ···················· | ···················· | ···················· | NNNNNNNNNNNNNNNNNNNN | NNNNN····NNNNNNNNNNN | 1 |
| M | 4691 hCoV-19/Wales/PHWC-47D134/2020\|EPI_ISL_586102\|2020 | ···················· | ···················· | ···················· | ···················· | NNNNNNNNNNNNNNNNNNNN | NNNNNNNNNNNNNNNNNNNN | 1 |
| M | 4692 hCoV-19/Wales/PHWC-47D143/2020\|EPI_ISL_586103\|2020 | ···················· | ···················· | ···················· | ···················· | NNNNNNNNNNNNNNNNNNNN | NNNNNNNNNNNNNNNNNNNN | 1 |
| M | 4694 hCoV-19/Wales/PHWC-47D301/2020\|EPI_ISL_586127\|2020 | ···················· | ···················· | ···················· | ···················· | NNNNNNNNNNNNNNNNNNNN | NNNN················ | 1 |
| M | 4699 hCoV-19/Wales/PHWC-47D3C5/2020\|EPI_ISL_586136\|2020 | ···················· | ···················· | ···················· | ···················· | NNNNNNNNNNNNNNNNNNNN | NNNNNNNNNNNNNNNNNNNN | 1 |
| M | 4701 hCoV-19/Wales/PHWC-47D3F2/2020\|EPI_ISL_586139\|2020 | ···················· | ···················· | ···················· | ···················· | NNNNNNNNNNNNNNNNNN·· | ···················· | 1 |
| M | 4704 hCoV-19/Wales/PHWC-47D43B/2020\|EPI_ISL_586142\|2020 | ···················· | ···················· | ···················· | ···················· | NNNNNNNNNNNNNNNNNNNN | ···················· | 1 |
| M | 4868 hCoV-19/Wales/PHWC-47D1F8/2020\|EPI_ISL_586111\|2020 | ···················· | ···················· | ···················· | ···················· | NNNNNNNNNNNNNNNNNNNN | NNNNNNNNNNNNNNNNNNNN | 1 |
| M | 4871 hCoV-19/Wales/PHWC-47D240/2020\|EPI_ISL_586116\|2020 | ···················· | ···················· | ···················· | ···················· | NNNNNNNNNNNNNNNNNNNN | NNNNNNNNNNNNNNNNNNNN | 1 |
| M | 4872 hCoV-19/Wales/PHWC-47D25F/2020\|EPI_ISL_586117\|2020 | ···················· | ···················· | ···················· | ···················· | NNNNNNNNNNNNNNNNNNNN | NNNNNNNNNNNNNNNNNNNN | 1 |
| M | 4873 hCoV-19/Wales/PHWC-47D26E/2020\|EPI_ISL_586118\|2020 | ···················· | ···················· | ···················· | ···················· | NNNNNNNNNNNNNNNNNNNN | ···················· | 1 |
| M | 4875 hCoV-19/Wales/PHWC-47D2B9/2020\|EPI_ISL_586122\|2020 | ···················· | ···················· | ···················· | ···················· | NNNNNNNNNNNNNNNNNNNN | NNNNNNNNNNNNNNNNNNNN | 1 |
| M | 4877 hCoV-19/Wales/PHWC-47D2E6/2020\|EPI_ISL_586125\|2020 | ···················· | ···················· | ···················· | ···················· | NNNNNNNNNNNNNNNNNNNN | NNNNNNNNNNNNNNNNNNNN | 1 |
| M | 5028 hCoV-19/Australia/VIC15296/2020\|EPI_ISL_593239\|202 | ···················· | ···················· | ···················· | ···················· | NNNNNNNNNNNNNNNN···· | ···················· | 1 |
| M | 5070 hCoV-19/Australia/VIC15393/2020\|EPI_ISL_593281\|202 | ···················· | ······A············· | ···················· | ···················· | NNNNNNNNNNNNNNNNNNNN | ···················· | 1 |
| M | 5073 hCoV-19/Australia/VIC15399/2020\|EPI_ISL_593284\|202 | ···················· | ···················· | ···················· | ···················· | NNNNNNNNNNNNNNNNNNNN | ···················· | 1 |
| M | 5083 hCoV-19/Australia/VIC15413/2020\|EPI_ISL_593294\|202 | ···················· | ···················· | ···················· | ···················· | ·NNNNNNNNNNN········ | ···················· | 1 |
| M | 5100 hCoV-19/England/BIRM-65607/2020\|EPI_ISL_594571\|202 | ···················· | ···················· | ···················· | ···················· | NNNNNNNNNNNNNNNNNNNN | NNNNNNNNNNNNNNNNNNNN | 1 |
| M | 5103 hCoV-19/England/BIRM-656CB/2020\|EPI_ISL_594576\|202 | ···················· | ···················· | ···················· | ···················· | NNNNNNNNNNNNNNNNNNNN | NNNNNNNNNNNNNNNNNNNN | 1 |
| M | 5106 hCoV-19/England/BIRM-65740/2020\|EPI_ISL_594579\|202 | ···················· | ···················· | ···················· | ···················· | NNNNNNNNNNNNNNNNNNNN | NNNNNNNNNNNNNNNNNNNN | 1 |
| M | 5107 hCoV-19/England/BIRM-6577D/2020\|EPI_ISL_594580\|202 | ···················· | ···················· | ···················· | ···················· | NNNNNNNNNNNNNNNNNNNN | NNNNNNNNNNNNNNNNNNNN | 1 |
| M | 5108 hCoV-19/England/BIRM-657E6/2020\|EPI_ISL_594584\|202 | ···················· | ···················· | ···················· | ···················· | NNNNNNNNNNNNNNNNNNNN | NNNNNNNNNNNNNNNNNNNN | 1 |
| M | 5112 hCoV-19/England/BIRM-658F2/2020\|EPI_ISL_594594\|202 | ···················· | ···················· | ···················· | ···················· | NNNNNNNNNNNNNNNNNNNN | NNNNNNNNNNNNNNNNNNNN | 1 |
| M | 5113 hCoV-19/England/BIRM-6591D/2020\|EPI_ISL_594595\|202 | ···················· | ···················· | ···················· | ···················· | NNNNNNNNNNNNNNNNNNNN | NNNNNNNNNNNNNNNNNNNN | 1 |
| M | 5115 hCoV-19/England/BIRM-659A4/2020\|EPI_ISL_594599\|202 | ···················· | ···················· | ···················· | ···················· | NNNNNNNNNNNNNNNNNNNN | NNNNNNNNNNNNNNNNNNNN | 1 |
| M | 5130 hCoV-19/England/CAMB-1B6378/2020\|EPI_ISL_584285\|20 | ···················· | ···················· | ···················· | ···················· | NNNNNNNNNNNNNNNNNNNN | NNNNNNNNNNNNNNNNNNNN | 1 |
| M | 5131 hCoV-19/England/CAMB-1B6554/2020\|EPI_ISL_584287\|20 | ···················· | ·A·················· | ···················· | ···················· | NNNNNNNNNNNNNNNNNNNN | NNNNNNNNNNNNNNNNNNNN | 1 |
| M | 5401 hCoV-19/England/CAMB-1B6606/2020\|EPI_ISL_584293\|20 | ···················· | ···················· | ···················· | ···················· | NNNNNNNNNNNNNNNNNNNN | NNNNNNNNNNNNNNNNNNNN | 1 |

| | | | | | | | | | |
|---|---|---|---|---|---|---|---|---|---|
| M | 5403 | hCoV-19/England/CAMB-1B66BB/2020\|EPI_ISL_584296\|20 | ···················· | ·A·················· | ···················· | ···················· | NNNNNNNNNNNNNNNNNNNN | NNNNNNNNNNNNNNNNNNNN | 1 |
| M | 5404 | hCoV-19/England/CAMB-1B66E8/2020\|EPI_ISL_584298\|20 | ···················· | ···················· | ···················· | ···················· | NNNNNNNNNNNNNNNNNNNN | NNNNNNNNNNNNNNNNNNNN | 1 |
| M | 5427 | hCoV-19/Scotland/QEUH-A00A53/2020\|EPI_ISL_585542\|2 | ···················· | ···················· | ···················· | ···················· | NNNNNNNNNNNNNNNNNNNN | NNNNNNNNNNNNNNNNNNNN | 1 |
| M | 5430 | hCoV-19/Scotland/QEUH-A00AEA/2020\|EPI_ISL_585547\|2 | ···················· | ···················· | ···················· | ···················· | NNNNNNNNNNNNNNNNNNNN | NNNNNNNNNNNNNNNNNNNN | 1 |
| M | 5431 | hCoV-19/Scotland/QEUH-A00B05/2020\|EPI_ISL_585549\|2 | ···················· | ···················· | ···················· | ···················· | NNNNNNNNNNNNNNNNNNNN | NNNNNNNNNNNNNNNNNNNN | 1 |
| M | 5435 | hCoV-19/Scotland/QEUH-A00B9C/2020\|EPI_ISL_585555\|2 | ···················· | ···················· | ···················· | ···················· | NNNNNNNNNNNNNNNNNNNN | NNNNNNNNNNNNNNNNNNNN | 1 |
| M | 5445 | hCoV-19/Scotland/QEUH-A00CA8/2020\|EPI_ISL_585568\|2 | ···················· | ···················· | ···················· | ···················· | NNNNNNNNNNNNNNNNNNNN | NNNNNNNNNNNNNNNNNNNN | 1 |
| M | 5454 | hCoV-19/Scotland/QEUH-A02B4F/2020\|EPI_ISL_585591\|2 | ···················· | ···················· | ···················· | ···················· | NNNNNNNNNNNNNNNNNNNN | NNNNNNNNNNNNNNNNNNNN | 1 |
| M | 5636 | hCoV-19/England/ALDP-9CC18C/2020\|EPI_ISL_584165\|20 | ···················· | ···················· | ···················· | ···················· | NNNNNNNNNNNNNNNNNNNN | NNNNNNNNNNNNNNNNNNNN | 1 |
| M | 5639 | hCoV-19/Scotland/EDB7939/2020\|EPI_ISL_585455\|2020- | ···················· | ···················· | ···················· | ···················· | NNNNNNNNNNNNNNNNNNNN | NNNNNNNNNNNNNNNNNNNN | 1 |
| M | 5713 | hCoV-19/env/Qingdao/IVDC-05-10/2020\|EPI_ISL_591274 | ···················· | ···················· | ···················· | ···················· | ··NNNNNNNNNNNNNNNNNN | NNNNNNNNNNNNNNNNNNNN | 1 |
| M | 5714 | hCoV-19/env/Qingdao/IVDC-06-10/2020\|EPI_ISL_591275 | ···················· | ···················· | ···················· | ···················· | NNNNNNNNNNNN········ | NNNNNNNNNNNNNNNNNNNN | 1 |
| M | 5728 | hCoV-19/Australia/VIC14435/2020\|EPI_ISL_592500\|202 | ···················· | ···················· | ···················· | ···················· | NNNNNNNNNNNNNNNNNNNN | NNNN················ | 1 |
| M | 5739 | hCoV-19/Australia/VIC14478/2020\|EPI_ISL_592512\|202 | ···················· | ···················· | ···················· | ···················· | NNNNNNNNNNNNNNNNNNNN | NNNNNNNNNNNNNNNNNNNN | 1 |
| M | 5745 | hCoV-19/Australia/VIC14491/2020\|EPI_ISL_592518\|202 | ···················· | ···················· | ···················· | ···················· | NNNNNNNNNNNNNNNNNNNN | ···················· | 1 |
| M | 5746 | hCoV-19/Australia/VIC14492/2020\|EPI_ISL_592519\|202 | ···················· | ···················· | ···················· | ···················· | NNNNNNNNNNNNNNNNNNNN | NNNNNNNNNNNNNNN····· | 1 |
| M | 5749 | hCoV-19/Australia/VIC14496/2020\|EPI_ISL_592522\|202 | ···················· | ···················· | ···················· | ···················· | ··NNNNNNNNNN········ | ···················· | 1 |
| M | 5752 | hCoV-19/Australia/VIC14501/2020\|EPI_ISL_592525\|202 | ···················· | ···················· | ···················· | ···················· | NNNNNNNNNNNNNNNNNNNN | NNNNNNNNNNNNNNNNNNNN | 1 |
| M | 5755 | hCoV-19/Australia/VIC14506/2020\|EPI_ISL_592528\|202 | ···················· | ···················· | ···················· | ···················· | NNNNNNNNNNNNNNNNN··· | ···················· | 1 |
| M | 5757 | hCoV-19/Australia/VIC14508/2020\|EPI_ISL_592530\|202 | ···················· | ···················· | ···················· | ···················· | NNNNNNNNNNNNNNNNNNNN | ··NN···NNNNNNNN····· | 1 |
| M | 5839 | hCoV-19/England/QEUH-A4CFC5/2020\|EPI_ISL_596076\|20 | ···················· | ···················· | ···················· | ···················· | NNNNNNNNNNNNNNNNNNNN | NNNNN·····N········· | 1 |
| M | 5882 | hCoV-19/Scotland/QEUH-A6372A/2020\|EPI_ISL_596138\|2 | ···················· | ···················· | ···················· | ···················· | NNNNNNNN·····N······ | ···················· | 1 |
| M | 5885 | hCoV-19/Scotland/QEUH-A63766/2020\|EPI_ISL_596142\|2 | ···················· | ···················· | ···················· | ···················· | NNNNNNNNNNNNNNNNNNNN | NNNNNNNNNNNNNNNNNNNN | 1 |
| M | 5893 | hCoV-19/Scotland/CVR4543/2020\|EPI_ISL_585402\|2020- | ···················· | ···················· | ···················· | ···················· | ·····N··N··········· | NN·····NN··········· | 1 |
| M | 5936 | hCoV-19/Scotland/EDB7988/2020\|EPI_ISL_585468\|2020- | ···················· | ···················· | ···················· | ···················· | NNNNNNNNNNNNNNNNNNNN | NNNNNNNNNNNNNNNNNNNN | 1 |
| M | 5937 | hCoV-19/Scotland/EDB7990/2020\|EPI_ISL_585469\|2020- | ···················· | ···················· | ···················· | ···················· | NNNNNNNNNNNNNNNNNNNN | NNNNNNNNNNNNNNNNNNNN | 1 |
| M | 6036 | hCoV-19/Australia/VIC14263/2020\|EPI_ISL_592424\|202 | ···················· | ···················· | ···················· | ···················· | NNNNNNNNNNNNNNNNNNNN | NN·················· | 1 |
| M | 6060 | hCoV-19/Australia/VIC14338/2020\|EPI_ISL_592457\|202 | ···················· | ···················· | ···················· | ···················· | NNNNNNNNNNNNNNNNNNNN | ···················· | 1 |
| M | 6064 | hCoV-19/Australia/VIC14349/2020\|EPI_ISL_592461\|202 | ···················· | ······A············· | ···················· | ···················· | NNNNNNNNNNNNNNNNNNNN | NNNNNNNNNNNNNNNNNNNN | 1 |
| M | 6117 | hCoV-19/Scotland/QEUH-A63890/2020\|EPI_ISL_596157\|2 | ···················· | ···················· | ···················· | ···················· | NNNNNNNNNNNNNNNNN··· | ···················· | 1 |
| M | 6159 | hCoV-19/Scotland/CVR4494/2020\|EPI_ISL_585357\|2020- | ···················· | ···················· | ···················· | ···················· | ···················· | NNNN················ | 1 |
| M | 6285 | hCoV-19/Australia/VIC14358/2020\|EPI_ISL_592466\|202 | ···················· | ···················· | ···················· | ···················· | NNNNNNNNNNNNNNNNNNNN | ···················· | 1 |
| M | 6292 | hCoV-19/Australia/VIC14373/2020\|EPI_ISL_592473\|202 | ···················· | ···················· | ···················· | ···················· | NNNNNNNNNNNNNNNNNNNN | ···················· | 1 |
| M | 6294 | hCoV-19/Australia/VIC14377/2020\|EPI_ISL_592475\|202 | ···················· | ···················· | ···················· | ···················· | ···················· | NNNNNNNNNNNNNNN····· | 1 |
| M | 6300 | hCoV-19/Australia/VIC14396/2020\|EPI_ISL_592481\|202 | ···················· | ···················· | ···················· | ···················· | NNNNNNNNNNNNNNNNNNNN | NNNNNNNNNNNNNNNNNNNN | 1 |
| M | 6301 | hCoV-19/Australia/VIC14397/2020\|EPI_ISL_592482\|202 | ···················· | ···················· | ···················· | ···················· | NNNNNNNNNNNNNNNNNNNN | ···················· | 1 |
| M | 6303 | hCoV-19/Australia/VIC14404/2020\|EPI_ISL_592484\|202 | ···················· | ···················· | ···················· | ···················· | NNNNNNNNNNNNNNNNNNNN | ···················· | 1 |
| M | 6315 | hCoV-19/Australia/VIC14422/2020\|EPI_ISL_592496\|202 | ···················· | ···················· | ···················· | ···················· | NNNNNNNNNNNNNNNNNNNN | ···················· | 1 |
| M | 6318 | hCoV-19/Australia/VIC14428/2020\|EPI_ISL_592499\|202 | ···················· | ···················· | ···················· | ···················· | NNNNNNNNNNNNNNNNNNNN | ···················· | 1 |
| M | 6384 | hCoV-19/Scotland/QEUH-A4C824/2020\|EPI_ISL_596004\|2 | ···················· | ···················· | ···················· | ···················· | NNNNNNNNNNNNNNNNNNNN | NNNNNNNNNNNNNNNNNNNN | 1 |
| M | 6388 | hCoV-19/Scotland/QEUH-A4C860/2020\|EPI_ISL_596008\|2 | ···················· | ···················· | ···················· | ···················· | NNNNNNNNNNNNNNNNNNNN | NNNNNNNNNNNNNNNNNNNN | 1 |
| M | 6397 | hCoV-19/Scotland/QEUH-A4CB2B/2020\|EPI_ISL_596021\|2 | ···················· | ···················· | ···················· | ···················· | NNNNNNNNN····NNNNNNN | NNN················· | 1 |
| M | 6398 | hCoV-19/England/QEUH-A4CB3A/2020\|EPI_ISL_596022\|20 | ···················· | ···················· | ···················· | ···················· | NNNNNNNNNNNNNNNNNNNN | NNNNNNNNNNNNNNNNNNNN | 1 |
| M | 6438 | hCoV-19/Scotland/CVR4442/2020\|EPI_ISL_585311\|2020- | ···················· | ···················· | ···················· | ···················· | ···················· | NN·····N·NN········· | 1 |
| M | 6461 | hCoV-19/Scotland/CVR4491/2020\|EPI_ISL_585354\|2020- | ···················· | ···················· | ···················· | ···················· | ···················· | NN·N···N············ | 1 |
| M | 6634 | hCoV-19/Scotland/QEUH-A4CC37/2020\|EPI_ISL_596035\|2 | ···················· | ···················· | ···················· | ···················· | NNNNNNNNNNNNNNNNNNNN | NNNNNNNNNNNNNNNNNNNN | 1 |
| M | 6642 | hCoV-19/Scotland/QEUH-A4CD25/2020\|EPI_ISL_596048\|2 | ···················· | ···················· | ···················· | ···················· | NNNNNNNNNNNNNNNNNNNN | ···················· | 1 |
| M | 6648 | hCoV-19/England/QEUH-A4CDE9/2020\|EPI_ISL_596057\|20 | ···················· | ···················· | ···················· | ···················· | NNNNNNNNNNNNNNNNNNNN | NNNNNNNNNNNNNNN····· | 1 |
| M | 6649 | hCoV-19/England/QEUH-A4CE04/2020\|EPI_ISL_596058\|20 | ···················· | ···················· | ···················· | ···················· | NNNNNNNNNNNNNNNNNNNN | NNNNNNNNNNNNNNNNNNNN | 1 |
| M | 6657 | hCoV-19/Northern Ireland/NIRE-108289/2020\|EPI_ISL_ | ···················· | ···················· | ···················· | ···················· | ···················· | NNNNNNNNNNNNNNN····· | 1 |
| M | 6658 | hCoV-19/Northern Ireland/NIRE-108298/2020\|EPI_ISL_ | ···················· | ···················· | ···················· | ···················· | NNNNNNNNNNNNNNNNNNNN | NNNNNNNNNNNNNNNNNNNN | 1 |
| M | 6659 | hCoV-19/Northern Ireland/NIRE-1082A7/2020\|EPI_ISL_ | ···················· | ···················· | ···················· | ···················· | ···················· | NNNNNNNNNNNNNNNN···· | 1 |
| M | 6715 | hCoV-19/Australia/VIC13697/2020\|EPI_ISL_592305\|202 | ···················· | ···················· | ···················· | ···················· | NNNNNNNNNNNNNNNNNNNN | NNNNNNNNNNNNNNNNN·NN | 1 |
| M | 6717 | hCoV-19/Australia/VIC13718/2020\|EPI_ISL_592307\|202 | ···················· | ···················· | ···················· | ···················· | NNNNNNNNNNNNNNNNNNNN | ···················· | 1 |
| M | 6718 | hCoV-19/Australia/VIC13721/2020\|EPI_ISL_592308\|202 | ···················· | ···················· | ···················· | ···················· | NNNNNNNNNNNNNNNNNNNN | NNNNNNNNNNNNNNNNNNNN | 1 |
| M | 6719 | hCoV-19/Australia/VIC13723/2020\|EPI_ISL_592309\|202 | ···················· | ···················· | ···················· | ···················· | NNNNNNNNNNNNNNNNNNNN | NNNNNNNNNNNNNNNNNNNN | 1 |
| M | 6737 | hCoV-19/Australia/VIC13839/2020\|EPI_ISL_592329\|202 | ···················· | ···················· | ···················· | ···················· | NNNNNNNNNNNNNNNNNNNN | NNNNNNNNNNNNNNNNNNNN | 1 |
| M | 6741 | hCoV-19/Australia/VIC13863/2020\|EPI_ISL_592333\|202 | ···················· | ···················· | ···················· | ···················· | NNNNNNNNNNNNNNNNNNNN | NNNNNNNNNNNNNNNNNNNN | 1 |
| M | 6744 | hCoV-19/Australia/VIC13874/2020\|EPI_ISL_592336\|202 | ···················· | ···················· | ···················· | ···················· | NNNNNNNNNNNNN······· | ···················· | 1 |
| M | 6749 | hCoV-19/Australia/VIC13944/2020\|EPI_ISL_592341\|202 | ···················· | ···················· | ···················· | ···················· | NNNN················ | ···················· | 1 |
| M | 6750 | hCoV-19/Australia/VIC14036/2020\|EPI_ISL_592342\|202 | ···················· | ···················· | ···················· | ···················· | NNNNNNNNNNNNNNNNNNNN | NNNNNNNNNNNNNNNNNNNN | 1 |
| M | 6751 | hCoV-19/Australia/VIC14066/2020\|EPI_ISL_592343\|202 | ···················· | ···················· | ···················· | ···················· | NNNN················ | ···················· | 1 |
| M | 6752 | hCoV-19/Australia/VIC14076/2020\|EPI_ISL_592344\|202 | ···················· | ···················· | ···················· | ···················· | NNNNNNNNNNNNNNNNNNNN | ···················· | 1 |
| M | 6755 | hCoV-19/Australia/VIC14122/2020\|EPI_ISL_592347\|202 | ···················· | ···················· | ···················· | ···················· | NNNNNNNNNNNNNNNNNNNN | ···················· | 1 |
| M | 6756 | hCoV-19/Australia/VIC14128/2020\|EPI_ISL_592348\|202 | ···················· | ···················· | ···················· | ···················· | ······R·NNNNNNNNNNNN | ···················· | 1 |
| M | 6768 | hCoV-19/Australia/VIC14161/2020\|EPI_ISL_592360\|202 | ···················· | ···················· | ···················· | ···················· | NNNNNNNNNNNNNNNNNNNN | ···················· | 1 |
| M | 6776 | hCoV-19/Australia/VIC14175/2020\|EPI_ISL_592368\|202 | ···················· | ···················· | ···················· | ···················· | NNNNNNNNNNNNNNNNNNNN | NNNNNNNNN··········· | 1 |
| M | 6777 | hCoV-19/Australia/VIC14176/2020\|EPI_ISL_592369\|202 | ···················· | ···················· | ···················· | ···················· | NNNNNNNNNNNNNNNNNNNN | NNNNNNNNNNNNNNNNNNNN | 1 |
| M | 6781 | hCoV-19/Australia/VIC14183/2020\|EPI_ISL_592373\|202 | ···················· | ···················· | ···················· | ···················· | NNNNNNNNNNNNN······· | ···················· | 1 |
| M | 6786 | hCoV-19/Australia/VIC14190/2020\|EPI_ISL_592378\|202 | ···················· | ···················· | ···················· | ···················· | NNNNNNNNNNNNNN······ | ···················· | 1 |
| M | 6788 | hCoV-19/Australia/VIC14193/2020\|EPI_ISL_592380\|202 | ···················· | ···················· | ···················· | ···················· | NNNNNNNNNNNNNNNNNNNN | ···················· | 1 |
| M | 6929 | hCoV-19/Northern Ireland/NIRE-FB369/2020\|EPI_ISL_5 | ···················· | ···················· | ···················· | ···················· | ·NNNNNNNN··········· | ···················· | 1 |
| M | 6931 | hCoV-19/Northern Ireland/NIRE-FB3A5/2020\|EPI_ISL_5 | ···················· | ···················· | ···················· | ···················· | NNNNNNNNNNNNNNNNNNNN | NNNN················ | 1 |
| M | 6934 | hCoV-19/Northern Ireland/NIRE-FB536/2020\|EPI_ISL_5 | ···················· | ···················· | ···················· | ···················· | NNNNNNNNNNNNNNNNNNNN | ···················· | 1 |
| M | 6936 | hCoV-19/Northern Ireland/NIRE-FB554/2020\|EPI_ISL_5 | ···················· | ···················· | ···················· | ···················· | ···················· | NNN········NNNN····· | 1 |
| M | 6940 | hCoV-19/Northern Ireland/NIRE-FB590/2020\|EPI_ISL_5 | ···················· | ···················· | ···················· | ···················· | ·NNNNNNNNNNNNNNNNNNN | NNN················· | 1 |
| M | 6947 | hCoV-19/Northern Ireland/NIRE-FB8D3/2020\|EPI_ISL_5 | ···················· | ···················· | ···················· | ···················· | NNNNNNNNNNNNNNNNNNNN | ···················· | 1 |
| M | 6955 | hCoV-19/Northern Ireland/NIRE-FB976/2020\|EPI_ISL_5 | ···················· | ···················· | ···················· | ···················· | NNNNNNNNNNNNNNNNNNNN | NNNNNNNNNNNNNNNNNNNN | 1 |
| M | 7101 | hCoV-19/DRC/16191/2020\|EPI_ISL_591086\|2020-06-03 | ···················· | ···················· | ···················· | ···················· | NNNNNNNNNNNNNNNNNNNN | NNNNNNNNNNNNNNNNNNNN | 1 |
| M | 7103 | hCoV-19/Australia/VIC14201/2020\|EPI_ISL_592385\|202 | ···················· | ···················· | ···················· | ···················· | NNNNNNNNNNNNNNNNNNNN | NNNNNNNNN··········· | 1 |

| | | | | | | | | | |
|---|---|---|---|---|---|---|---|---|---|
| M | 7107 | hCoV-19/Australia/VIC14207/2020\|EPI_ISL_592389\|202 | ···················· | ···················· | ···················· | ···················· | NNNNNNNNNNNNNNNNNNNN | NNNN················ | 1 |
| M | 7174 | hCoV-19/England/LIVE-A0CC2/2020\|EPI_ISL_584615\|202 | ···················· | ···················· | ···················· | ···················· | NNNNNNNNNNNNNNNNNNNN | NNNNNNNNNNNNNNNNNNNN | 1 |
| M | 7177 | hCoV-19/England/LIVE-A4578/2020\|EPI_ISL_584619\|202 | ···················· | ···················· | ···················· | ···················· | ·······NNNNNNNNNNNNN | NNNNNNNNNNNNNNNNNNNN | 1 |
| M | 7178 | hCoV-19/England/LIVE-A45B4/2020\|EPI_ISL_584620\|202 | ···················· | ···················· | ···················· | ···················· | ·······NNNNNNNNNNNNN | NNNNNNNNNNNNNNNNNNNN | 1 |
| M | 7184 | hCoV-19/England/LOND-124D9BC/2020\|EPI_ISL_584629\|2 | ···················· | ···················· | ···················· | ···················· | NNNNNNNNNNNNNNNNNNNN | NNNNNNNNNNNNNNNNNNNN | 1 |
| M | 7244 | hCoV-19/Wales/PHWC-16F36F/2020\|EPI_ISL_585952\|2020 | ···················· | ···················· | ···················· | ···················· | NNNNNNNNNNNNNNNNNNNN | NNNNNNNNNNNNNNNNNNNN | 1 |
| M | 7618 | hCoV-19/Australia/VIC14064/2020\|EPI_ISL_591794\|202 | ···················· | ···················· | ···················· | ···················· | NNNNNNNNNNNN········ | ···················· | 1 |
| M | 7626 | hCoV-19/Australia/VIC13864/2020\|EPI_ISL_591802\|202 | ···················· | ···················· | ···················· | ···················· | NNNNNNNNNNNNNNNNNNNN | NNNNNNNNNNNNNNNNNNNN | 1 |
| M | 7630 | hCoV-19/Australia/VIC14326/2020\|EPI_ISL_591806\|202 | ···················· | ···················· | ···················· | ···················· | NNNNNNNNNNNNNNNNNNNN | ···················· | 1 |
| M | 7631 | hCoV-19/Australia/VIC14366/2020\|EPI_ISL_591807\|202 | ···················· | ···················· | ···················· | ···················· | NNNNNNNNNNNNNNNNNNNN | ···················· | 1 |
| M | 7633 | hCoV-19/Australia/VIC11664/2020\|EPI_ISL_591831\|202 | ···················· | ···················· | ···················· | ···················· | ······R····NNNNNNNNN | NN·················· | 1 |
| M | 7634 | hCoV-19/Australia/VIC11668/2020\|EPI_ISL_591832\|202 | ···················· | ···················· | ···················· | ···················· | NNNN················ | ···················· | 1 |
| M | 7641 | hCoV-19/Australia/VIC11714/2020\|EPI_ISL_591839\|202 | ···················· | ···················· | ···················· | ···················· | NNNNNNNNNNNNNNNNNNNN | NNNNNNNNNNNNNNNNNNNN | 1 |
| M | 7643 | hCoV-19/Australia/VIC11737/2020\|EPI_ISL_591841\|202 | ···················· | ···················· | ···················· | ···················· | NNNNNNNNNNNNNNNNNNNN | NNNNN··············· | 1 |
| M | 7644 | hCoV-19/Australia/VIC11748/2020\|EPI_ISL_591842\|202 | ···················· | ···················· | ···················· | ···················· | ······R··NNNNNNNNNNN | NNNNNNNNNNNNNNNNNNNN | 1 |
| M | 7645 | hCoV-19/Australia/VIC11776/2020\|EPI_ISL_591847\|202 | ···················· | ···················· | ···················· | ···················· | NNNNNNNNNNNNNNNNNNNN | NNNNNNNNNNNNNNNNNNNN | 1 |
| M | 7680 | hCoV-19/Wales/PHWC-47E072/2020\|EPI_ISL_595399\|2020 | ···················· | ···················· | ···················· | ···················· | NNNNNNNNNNN········· | ···················· | 1 |
| M | 7686 | hCoV-19/England/LOND-124DD47/2020\|EPI_ISL_584648\|2 | ···················· | ···················· | ···················· | ···················· | NNNNNNNNNNNNNNNNNNNN | NNNNNNNNNNNNNNNNNNNN | 1 |
| M | 7687 | hCoV-19/England/LOND-124DDB0/2020\|EPI_ISL_584650\|2 | ···················· | ···················· | ···················· | ···················· | NNNNNNNNNNNNNNNNNNNN | NNNNNNNNNNNNNNNNNNNN | 1 |
| M | 7689 | hCoV-19/England/LOND-124DE44/2020\|EPI_ISL_584652\|2 | ···················· | ···················· | ···················· | ···················· | NNNNNNNNNNNNNNNNNNNN | NNNNNNNNNNNNNNNNNNNN | 1 |
| M | 7704 | hCoV-19/England/NORT-29CDF1/2020\|EPI_ISL_584681\|20 | ···················· | ···················· | ···················· | ···················· | NNNNNNNNNNNNNNNNNNNN | NNNNNNNNNNNNNNNNNNNN | 1 |
| M | 7727 | hCoV-19/Wales/PHWC-16EA2F/2020\|EPI_ISL_585807\|2020 | ···················· | ···················· | ···················· | ···················· | NNNNNNNNNNNNNNNNNNNN | ···················· | 1 |
| M | 7731 | hCoV-19/Wales/PHWC-16F4B7/2020\|EPI_ISL_585973\|2020 | ···················· | ···················· | ···················· | ···················· | NNNNNNNNNNNNNNNNNNNN | NNNNNNNNNNNNNNNNNNNN | 1 |
| M | 7735 | hCoV-19/Wales/PHWC-16F4F3/2020\|EPI_ISL_585977\|2020 | ···················· | ···················· | ···················· | ···················· | NNNNNNNNNNNNNNNNNNNN | ···················· | 1 |
| M | 7841 | hCoV-19/Australia/VIC12904/2020\|EPI_ISL_591811\|202 | ···················· | ···················· | ···················· | ···················· | NNN···NNNNNN········ | NNNNNNNNNNNNNNN····· | 1 |
| M | 7845 | hCoV-19/Australia/VIC11606/2020\|EPI_ISL_591826\|202 | ···················· | ···················· | ···················· | ···················· | NNNNNNNNNNNNNNNNNNNN | NNNNNNNNNNNNNNNNNNNN | 1 |
| M | 7846 | hCoV-19/Australia/VIC11610/2020\|EPI_ISL_591827\|202 | ···················· | ···················· | ···················· | ···················· | NNNNNNNNNNNNNNNNNNNN | NNNNNNNNNNNNNNNNNNNN | 1 |
| M | 7848 | hCoV-19/Australia/VIC11653/2020\|EPI_ISL_591829\|202 | ···················· | ···················· | ···················· | ···················· | NNNNNNNNNNNNNNNNNNNN | NNNNNNNNNNNNNNN····· | 1 |
| M | 7905 | hCoV-19/Wales/PHWC-47DBAF/2020\|EPI_ISL_595335\|2020 | ···················· | ···················· | ···················· | ···················· | NNNNNNNNNNNNNNNNNNNN | ···················· | 1 |
| M | 7916 | hCoV-19/Wales/PHWC-47DC8E/2020\|EPI_ISL_595347\|2020 | ···················· | ···················· | ···················· | ···················· | NNNNNNNNNNNNNNNNNNNN | NNNNNNNNNNNNNNNNNNNN | 1 |
| M | 7919 | hCoV-19/Wales/PHWC-47DCD9/2020\|EPI_ISL_595351\|2020 | ···················· | ···················· | ···················· | ···················· | NNNNNNNNNNNNNNNNNNNN | ···················N | 1 |
| M | 7925 | hCoV-19/Wales/PHWC-47DD7C/2020\|EPI_ISL_595360\|2020 | ···················· | ···················· | ···················· | ···················· | NNNNNNNNNNNNNNNN···· | NNNNNNNNN···N······· | 1 |
| M | 7930 | hCoV-19/Wales/PHWC-47DE2E/2020\|EPI_ISL_595367\|2020 | ···················· | ···················· | ···················· | ···················· | NNNNNNNNNNNNNNNNNNNN | ···················· | 1 |
| M | 7931 | hCoV-19/Wales/PHWC-47DE3D/2020\|EPI_ISL_595368\|2020 | ···················· | ···················· | ···················· | ···················· | NNNNNNNNNNNNNN······ | ···················· | 1 |
| M | 8148 | hCoV-19/Australia/VIC14661/2020\|EPI_ISL_592940\|202 | ···················· | ···················· | ···················· | ···················· | NNNNNNNNNNN········· | ···················· | 1 |
| M | 8150 | hCoV-19/Australia/VIC14663/2020\|EPI_ISL_592942\|202 | ···················· | ······A············· | ···················· | ···················· | ···················· | NNNNNNNNNNNNNNN····· | 1 |
| M | 8155 | hCoV-19/Australia/VIC14671/2020\|EPI_ISL_592947\|202 | ···················· | ···················· | ···················· | ···················· | NNNNNNNNNNNNNNNNNNNN | NNNNNNNNNNNNNNNNNNNN | 1 |
| M | 8167 | hCoV-19/Australia/VIC14703/2020\|EPI_ISL_592959\|202 | ···················· | ···················· | ···················· | ···················· | NNNNNNNNNNNNNNNNNNNN | NNNNNNNNNNNNNNNNNNNN | 1 |
| M | 8171 | hCoV-19/Australia/VIC14709/2020\|EPI_ISL_592963\|202 | ···················· | ···················· | ···················· | ···················· | NNNNNNNNNNNNNNNNNNNN | NNNNNM····NNNNNNNNNN | 1 |
| M | 8172 | hCoV-19/Australia/VIC14712/2020\|EPI_ISL_592964\|202 | ···················· | ···················· | ···················· | ···················· | NNNNNNNNNNNNNNNNNNNN | NNNNNNNNNNNNNNNNNNNN | 1 |
| M | 8175 | hCoV-19/Australia/VIC14717/2020\|EPI_ISL_592967\|202 | ···················· | ···················· | ···················· | ···················· | NNNNNNNNNNNNNNNNNNNN | NNNNNNNNNNNNNNN····· | 1 |
| M | 8181 | hCoV-19/Australia/VIC14729/2020\|EPI_ISL_592973\|202 | ···················· | ···················· | ···················· | ···················· | NNNNNNNNNNNN········ | ···················· | 1 |
| M | 8231 | hCoV-19/England/CWAR-24FE153/2020\|EPI_ISL_584578\|2 | ···················· | ···················· | ···················· | ···················· | NNNNNNNNNNNNNNNNNNNN | NNNNNNNNNNNNNNNNNNNN | 1 |
| M | 8241 | hCoV-19/Wales/PHWC-16ED62/2020\|EPI_ISL_585858\|2020 | ···················· | ···················· | ···················· | ···················· | NNNN··NNNNNNNNNNNNNN | ···················· | 1 |
| M | 8264 | hCoV-19/Wales/PHWC-16EF11/2020\|EPI_ISL_585885\|2020 | ···················· | ···················· | ···················· | ···················· | NNNNNNNNNNNNNNNNNNNN | NNNNN··············· | 1 |
| M | 8368 | hCoV-19/Australia/VIC14741/2020\|EPI_ISL_592980\|202 | ···················· | ···················· | ···················· | ···················· | NNN···NNNNN········· | ···················· | 1 |
| M | 8379 | hCoV-19/Australia/VIC14779/2020\|EPI_ISL_592996\|202 | ···················· | ···················· | ···················· | ···················· | NNNNNNNNNNNNNNNNNNNN | NNNNNNNNNNNNNNNNNNNN | 1 |
| M | 8388 | hCoV-19/England/NORW-F0CC3/2020\|EPI_ISL_595212\|202 | ···················· | ···················· | ···················· | ···················· | ···················· | NNNNN··············· | 1 |
| M | 8389 | hCoV-19/England/NORW-F0CE1/2020\|EPI_ISL_595213\|202 | ···················· | ···················· | ···················· | ···················· | ···················· | NNNNNNNNNNNNNNN····· | 1 |
| M | 8393 | hCoV-19/England/NORW-F1390/2020\|EPI_ISL_595227\|202 | ···················· | ···················· | ···················· | ···················· | NNNNNNNNNNNNNN······ | NNNNNNNNNNNNNNN····· | 1 |
| M | 8395 | hCoV-19/England/NORW-F1460/2020\|EPI_ISL_595229\|202 | ···················· | ···················· | ···················· | ···················· | NNNNNNNNNNNNNNNNNNNN | NNNNNNNNNNNNNNNNNNNN | 1 |
| M | 8396 | hCoV-19/England/NORW-F14AC/2020\|EPI_ISL_595231\|202 | ···················· | ···················· | ···················· | ···················· | NNNNNNNNNNNNNNNNNNNN | NNNNNNNNNNNNNNNNNNNN | 1 |
| M | 8449 | hCoV-19/Wales/PHWC-16E306/2020\|EPI_ISL_585700\|2020 | ···················· | ···················· | ···················· | ···················· | NNNNNNNNNNNNNNNNNNNN | ···················· | 1 |
| M | 8451 | hCoV-19/Wales/PHWC-16E333/2020\|EPI_ISL_585702\|2020 | ···················· | ···················· | ···················· | ···················· | NNNNNNNNNNNNNNNNNNNN | ···················· | 1 |
| M | 8453 | hCoV-19/Wales/PHWC-16E360/2020\|EPI_ISL_585705\|2020 | ···················· | ···················· | ···················· | ···················· | NNNNNNNNNNNNNNNNNNNN | NNNNNNNNNNNNNNNNNNNN | 1 |
| M | 8457 | hCoV-19/Wales/PHWC-16E3BB/2020\|EPI_ISL_585709\|2020 | ···················· | ···················· | ···················· | ···················· | NNNNNNNNNNNNNNNNNNNN | ···················· | 1 |
| M | 8458 | hCoV-19/Wales/PHWC-16E3CA/2020\|EPI_ISL_585710\|2020 | ···················· | ···················· | ···················· | ···················· | NNNNNNNNNNNNNNNNNNNN | NNNNNNNNNNNNNNNNNNNN | 1 |
| M | 8460 | hCoV-19/Wales/PHWC-16E3E8/2020\|EPI_ISL_585712\|2020 | ···················· | ···················· | ···················· | ···················· | NNNNNNNNNNNNNNNNNNNN | NNNNNNNNNNNNNNNNNNNN | 1 |
| M | 8463 | hCoV-19/Wales/PHWC-16E46D/2020\|EPI_ISL_585719\|2020 | ···················· | ···················· | ···················· | ···················· | NNNNNNNNNNNNNNNNNNNN | ···················· | 1 |
| M | 8464 | hCoV-19/Wales/PHWC-16E47C/2020\|EPI_ISL_585720\|2020 | ···················· | ···················· | ···················· | ···················· | NNNNNNNNNNNNNNNNNNNN | ···················· | 1 |
| M | 8465 | hCoV-19/Wales/PHWC-16E48B/2020\|EPI_ISL_585721\|2020 | ···················· | ···················· | ···················· | ···················· | NNNNNNNNNNNNNNNNNNNN | NNNNN··············· | 1 |
| M | 8467 | hCoV-19/Wales/PHWC-16E4A9/2020\|EPI_ISL_585723\|2020 | ···················· | ···················· | ···················· | ···················· | NNNNNNNNNNNNNNNNNNNN | ···················· | 1 |
| M | 8469 | hCoV-19/Wales/PHWC-16E4D6/2020\|EPI_ISL_585726\|2020 | ···················· | ···················· | ···················· | ···················· | NNNNNNNNNNNNNNNNNNNN | ···················· | 1 |
| M | 8647 | hCoV-19/Australia/VIC14819/2020\|EPI_ISL_592827\|202 | ···················· | ···················· | ···················· | ···················· | NNNNNNNNNNNNNNNNNNNN | ···················· | 1 |
| M | 8648 | hCoV-19/Australia/VIC15071/2020\|EPI_ISL_592828\|202 | ···················· | ···················· | ···················· | ···················· | NNNNNNNNNNNNNNNNNNNN | ···················· | 1 |
| M | 8652 | hCoV-19/Australia/VIC15824/2020\|EPI_ISL_592832\|202 | ···················· | ···················· | ···················· | ···················· | NNNNNNNNNNNNNNNNNNNN | ···················· | 1 |
| M | 8661 | hCoV-19/Australia/VIC14904/2020\|EPI_ISL_592841\|202 | ···················· | ···················· | ···················· | ···················· | NNNNNNNNNNNNNNNNNNNN | ···················· | 1 |
| M | 8674 | hCoV-19/Australia/VIC15644/2020\|EPI_ISL_592854\|202 | ···················· | ···················· | ···················· | ···················· | NNNNNNNNNNNNNN······ | ···················· | 1 |
| M | 8675 | hCoV-19/Australia/VIC15749/2020\|EPI_ISL_592855\|202 | ···················· | ···················· | ···················· | ···················· | NNNNNNNNNNNNNNNNNNNN | NNNN··········NNNNNN | 1 |
| M | 8679 | hCoV-19/Australia/VIC14512/2020\|EPI_ISL_592868\|202 | ···················· | ···················· | ···················· | ···················· | NNNNNNNNNNNNNNNNNNNN | ···················· | 1 |
| M | 8684 | hCoV-19/Australia/VIC14523/2020\|EPI_ISL_592876\|202 | ···················· | ···················· | ···················· | ···················· | NNNNNNNNNNNNNNNNNNNN | NNNN················ | 1 |
| M | 8688 | hCoV-19/Australia/VIC14543/2020\|EPI_ISL_592883\|202 | ···················· | ···················· | ···················· | ···················· | NNNNNNNNNNNNNNNNNNNN | ···················· | 1 |
| M | 8690 | hCoV-19/Australia/VIC14545/2020\|EPI_ISL_592885\|202 | ···················· | ···················· | ···················· | ···················· | NNNNNNNNNNNNNNNNNNNN | ···················· | 1 |
| M | 8691 | hCoV-19/Australia/VIC14546/2020\|EPI_ISL_592886\|202 | ···················· | ···················· | ···················· | ···················· | NNNNNNNNNNNNNNNNNNNN | ···················· | 1 |
| M | 8692 | hCoV-19/Australia/VIC14547/2020\|EPI_ISL_592887\|202 | ···················· | ···················· | ···················· | ···················· | NNNNNNNNNNNNNNNNNNNN | ···················· | 1 |
| M | 8693 | hCoV-19/Australia/VIC14549/2020\|EPI_ISL_592888\|202 | ···················· | ···················· | ···················· | ···················· | NNNNNNNNNNNNNNNNNNNN | ··············N····· | 1 |
| M | 8694 | hCoV-19/Australia/VIC14551/2020\|EPI_ISL_592889\|202 | ···················· | ···················· | ···················· | ···················· | NNNNNNNNNNNNNNNNNNNN | NNNNNNNNNNNNNNNNNNNN | 1 |
| M | 8697 | hCoV-19/Australia/VIC14557/2020\|EPI_ISL_592892\|202 | ···················· | ···················· | ···················· | ···················· | NNNNN··············· | ···················· | 1 |
| M | 8701 | hCoV-19/Australia/VIC14565/2020\|EPI_ISL_592896\|202 | ···················· | ···················· | ···················· | ···················· | NNNNNNNNNNNNNNNNNNNN | ···················· | 1 |
| M | 8742 | hCoV-19/England/CWAR-24FDA1D/2020\|EPI_ISL_584466\|2 | ···················· | ···················A | ···················· | ···················· | NNNNNNNNNNNNNNNNNNNN | NNNNNNNNNNNNNNNNNNNN | 1 |

| | | | | | | | | | |
|---|---|---|---|---|---|---|---|---|---|
| M | 8752 | hCoV-19/England/CWAR-24FDAB3/2020\|EPI_ISL_584476\|2 | ···················· | ···················· | ···················· | ···················· | NNNNNNNNNNNNNNNNNNNN | NNNNNNNNNNNNNNNNNNNN | 1 |
| M | 8770 | hCoV-19/Wales/PHWC-16E55B/2020\|EPI_ISL_585733\|2020 | ···················· | ···················· | ···················· | ···················· | NNNNNNNNNNNNNNNNNNNN | NNNNNNNNNNNNNNNNNNNN | 1 |
| M | 8773 | hCoV-19/Wales/PHWC-16E588/2020\|EPI_ISL_585736\|2020 | ···················· | ···················· | ···················· | ···················· | NNNNNNNNNNNNNNNNNNNN | ···················· | 1 |
| M | 8775 | hCoV-19/Wales/PHWC-16E5A6/2020\|EPI_ISL_585738\|2020 | ···················· | ···················· | ···················· | ···················· | NNNNNNNNNNNNNNNNNNNN | ···················· | 1 |
| M | 8777 | hCoV-19/Wales/PHWC-16E5D3/2020\|EPI_ISL_585741\|2020 | ···················· | ···················· | ···················· | ···················· | NNNNNNNNNNN·NNNNNNNN | ···················· | 1 |
| M | 8782 | hCoV-19/Wales/PHWC-16E649/2020\|EPI_ISL_585747\|2020 | ···················· | ···················· | ···················· | ···················· | NNNNNNNNNNNNNNNNNNNN | NNNNNNNNNNNNNNNNNNNN | 1 |
| M | 8783 | hCoV-19/Wales/PHWC-16E667/2020\|EPI_ISL_585749\|2020 | ···················· | ···················· | ···················· | ···················· | NNNNNNNNNNNNNNNNNNNN | NNNNNNNNNNNNNNNNNNNN | 1 |
| M | 8784 | hCoV-19/Wales/PHWC-16E676/2020\|EPI_ISL_585750\|2020 | ···················· | ···················· | ···················· | ···················· | NNNNNNNNNNNNNNNNNNNN | ···················· | 1 |
| M | 8785 | hCoV-19/Wales/PHWC-16E694/2020\|EPI_ISL_585752\|2020 | ···················· | ···················· | ···················· | ···················· | NNNNNNNNNNNNNNNNNNNN | ···················· | 1 |
| M | 8788 | hCoV-19/Wales/PHWC-16E6D0/2020\|EPI_ISL_585756\|2020 | ···················· | ···················· | ···················· | ···················· | NNNNNNNNNNNNNNNNNNNN | NNNNNNNNNNNNNNNNNNNN | 1 |
| M | 8789 | hCoV-19/Wales/PHWC-16E6FE/2020\|EPI_ISL_585758\|2020 | ···················· | ···················· | ···················· | ···················· | NNNNNNNNNNNNNNNNNNNN | NNNNNNNNNNNNNNNNNNNN | 1 |
| M | 8790 | hCoV-19/Wales/PHWC-16E719/2020\|EPI_ISL_585760\|2020 | ···················· | ···················· | ···················· | ···················· | NNNNNNNNNNNNNNNNNNNN | ···················· | 1 |
| M | 8792 | hCoV-19/Wales/PHWC-16E773/2020\|EPI_ISL_585765\|2020 | ···················· | ···················· | ···················· | ···················· | NNNNNNNNNNNNNNNNNNNN | NNNNNNNNNNNNNNNNNNNN | 1 |
| M | 8799 | hCoV-19/Wales/PHWC-16E825/2020\|EPI_ISL_585775\|2020 | ···················· | ···················· | ···················· | ···················· | NNNNNNNNNNNNNNNNNNNN | NNNNNNNNNNNNNNNNNNNN | 1 |
| M | 8801 | hCoV-19/Wales/PHWC-16E843/2020\|EPI_ISL_585777\|2020 | ···················· | ···················· | ···················· | ···················· | NNNNNNNNNNNNNNNNNNNN | ···················· | 1 |
| M | 8813 | hCoV-19/Wales/PHWC-16E913/2020\|EPI_ISL_585790\|2020 | ···················· | ···················· | ···················· | ···················· | NNNNNNNNNNNNNNNNNNNN | NNNNNNNNNNNNNNN····· | 1 |
| M | 8868 | hCoV-19/Australia/VIC14516/2020\|EPI_ISL_592872\|202 | ···················· | ···················· | ···················· | ···················· | NNNNNNNNNNNN·NNNNNNN | NNNNNNNNNNNNNNN····· | 1 |
| M | 8872 | hCoV-19/Australia/VIC14570/2020\|EPI_ISL_592900\|202 | ···················· | ···················· | ···················· | ···················· | NNNN················ | ···················· | 1 |
| M | 8877 | hCoV-19/Australia/VIC14585/2020\|EPI_ISL_592905\|202 | ···················· | ···················· | ···················· | ···················· | NNNNNNNNNNNNNNNNNNNN | ···················· | 1 |
| M | 8879 | hCoV-19/Australia/VIC14590/2020\|EPI_ISL_592907\|202 | ···················· | ···················· | ···················· | ···················· | NNNNNNNNNNNNNNNNNNNN | NNNNNNNNNNNNNNNNNNNN | 1 |
| M | 8880 | hCoV-19/Australia/VIC14592/2020\|EPI_ISL_592908\|202 | ···················· | ···················· | ···················· | ···················· | NNNNNNNNNNNNNNNNNNNN | NNNNNNNNNNNNNNNNNNNN | 1 |
| M | 8881 | hCoV-19/Australia/VIC14593/2020\|EPI_ISL_592909\|202 | ···················· | ···················· | ···················· | ···················· | NNNNNNNNNNNNNNNN···· | ···················· | 1 |
| M | 8886 | hCoV-19/Australia/VIC14610/2020\|EPI_ISL_592914\|202 | ···················· | ···················· | ···················· | ···················· | NNNNNNNNNNNNNNNNNNNN | ···················· | 1 |
| M | 8888 | hCoV-19/Australia/VIC14613/2020\|EPI_ISL_592916\|202 | ···················· | ···················· | ···················· | ···················· | NNNNNNNNNNNNNNNNNNNN | ···················· | 1 |
| M | 8890 | hCoV-19/Australia/VIC14620/2020\|EPI_ISL_592918\|202 | ···················· | ···················· | ···················· | ···················· | NNNNNNNNNNNNNNNNNNNN | NNNNNNNNNNNNNNNNNNNN | 1 |
| M | 8892 | hCoV-19/Australia/VIC14624/2020\|EPI_ISL_592920\|202 | ···················· | ···················· | ···················· | ···················· | NNNNNNNNNNNNNNNNNNNN | ···················· | 1 |
| M | 8894 | hCoV-19/Australia/VIC14626/2020\|EPI_ISL_592922\|202 | ···················· | ·····A·············· | ···················· | ···················· | NNNNNNNNNNNNNNNNNNNN | NNNNNNNNNNNNNNN····· | 1 |
| M | 8901 | hCoV-19/Australia/VIC14640/2020\|EPI_ISL_592929\|202 | ···················· | ···················· | ···················· | ···················· | NNNNNNNNNNNNNNNNNNNN | NNNNNNNNNNNNNNNNNNNN | 1 |
| M | 8905 | hCoV-19/Australia/VIC14649/2020\|EPI_ISL_592933\|202 | ···················· | ···················· | ···················· | ···················· | NNNNNNNNNNNNNNNNNNNN | ···················· | 1 |
| M | 8908 | hCoV-19/Australia/VIC14655/2020\|EPI_ISL_592936\|202 | ···················· | ···················· | ···················· | ···················· | NNNNNNNNNNNNNNNNNNNN | ···················· | 1 |
| M | 8932 | hCoV-19/England/MILK-A5605C/2020\|EPI_ISL_595130\|20 | ···················· | ···················· | ···················· | ···················· | NNNNNNNNNNNNNNNNNNNN | NNNNNNNNNNN········· | 1 |
| M | 8948 | hCoV-19/England/MILK-A5639F/2020\|EPI_ISL_595151\|20 | ···················· | ···················· | ···················· | ···················· | NNNNNNNNNNNNNNNNNNNN | NNNNNNNNNNNNNNNNNNNN | 1 |
| M | 8952 | hCoV-19/England/NORW-EFDB5/2020\|EPI_ISL_595159\|202 | ···················· | ···················· | ···················· | ···················· | NNNNNNNNNNNNNNNNNNNN | NNNNNNNNNNN········· | 1 |
| M | 8953 | hCoV-19/England/NORW-EFE0D/2020\|EPI_ISL_595161\|202 | ···················· | ···················· | ···················· | ···················· | NNNNNNNNNNNNNNNNNNNN | NNNNNNNNNNNNNNNNNNNN | 1 |
| M | 8955 | hCoV-19/England/NORW-EFFEC/2020\|EPI_ISL_595169\|202 | ···················· | ···················· | ···················· | ···················· | NNNNNNNNNNNNNNNNNNNN | NNNNNNNNNNNNNNNNNNNN | 1 |
| M | 8956 | hCoV-19/England/NORW-F0021/2020\|EPI_ISL_595172\|202 | ···················· | ···················· | ···················· | ···················· | NNNNNNNNNNNNNNNNNNNN | NNNNN··············· | 1 |
| M | 8957 | hCoV-19/England/NORW-F013D/2020\|EPI_ISL_595177\|202 | ···················· | ···················· | ···················· | ···················· | ···················· | NNNNN··············· | 1 |
| M | 8958 | hCoV-19/England/NORW-F020D/2020\|EPI_ISL_595179\|202 | ···················· | ···················· | ···················· | ···················· | NNNNNNNNNNNNNNNNNNNN | NNNNN··············· | 1 |
| M | 8959 | hCoV-19/England/NORW-F05AA/2020\|EPI_ISL_595184\|202 | ···················· | ···················· | ···················· | ···················· | ···················· | NNNNN··············· | 1 |
| M | 8960 | hCoV-19/England/NORW-F0768/2020\|EPI_ISL_595190\|202 | ···················· | ···················· | ···················· | ···················· | NNNNNNNNNNNNNNNNNNNN | NNNNNNNNNNN········· | 1 |
| M | 8961 | hCoV-19/Scotland/QEUH-A0C1BC/2020\|EPI_ISL_585603\|2 | ···················· | ···················· | ···················· | ···················· | NNN················· | NNNNNNNNNNNNNNNNN··N | 1 |
| M | 9219 | hCoV-19/England/CAMB-1B677C/2020\|EPI_ISL_584304\|20 | ···················· | ···················· | ···················· | ···················· | NNNNNNNNNNNNNNNNNNNN | NNNNNNNNNNNNNNNNNNNN | 1 |
| M | 9220 | hCoV-19/England/CAMB-1B679A/2020\|EPI_ISL_584305\|20 | ···················· | ···················· | ···················· | ···················· | NNNNNNNNNNNNNNNNNNNN | NNNNNNNNNNNNNNNNNNNN | 1 |
| M | 9221 | hCoV-19/England/CAMB-1B67D6/2020\|EPI_ISL_584307\|20 | ···················· | ···················· | ···················· | ···················· | NNNNNNNNNNNNNNNNNNNN | NNNNNNNNNNNNNNNNNNNN | 1 |
| M | 9222 | hCoV-19/England/CAMB-1B6897/2020\|EPI_ISL_584310\|20 | ···················· | ···················· | ···················· | ···················· | NNNNNNNNNNNNNNNNNNNN | NNNNNNNNNNNNNNNNNNNN | 1 |
| M | 9226 | hCoV-19/England/CAMB-1B68F1/2020\|EPI_ISL_584314\|20 | ···················· | ···················· | ···················· | ···················· | NNNNNNNNNNNNNNNNNNNN | NNNNNNNNNNNNNNNNNNNN | 1 |
| M | 9227 | hCoV-19/England/CAMB-1B690D/2020\|EPI_ISL_584315\|20 | ···················· | ···················· | ···················· | ···················· | NNNNNNNNNNNNNNNNNNNN | NNNNNNNNNNNNNNNNNNNN | 1 |
| M | 9229 | hCoV-19/England/CAMB-1B692B/2020\|EPI_ISL_584317\|20 | ···················· | ···················· | ···················· | ···················· | NNNNNNNNNNNNNNNNNNNN | NNNNNNNNNNNNNNNNNNNN | 1 |
| M | 9231 | hCoV-19/England/CAMB-1B69A3/2020\|EPI_ISL_584320\|20 | ···················· | ···················· | ···················· | ···················· | NNNNNNNNNNNNNNNNNNNN | NNNNNNNNNNNNNNNNNNNN | 1 |
| M | 9232 | hCoV-19/England/CAMB-1B69D0/2020\|EPI_ISL_584321\|20 | ···················· | ·A·················· | ···················· | ···················· | NNNNNNNNNNNNNNNNNNNN | NNNNNNNNNNNNNNNNNNNN | 1 |
| M | 9265 | hCoV-19/Scotland/QEUH-A0CA48/2020\|EPI_ISL_585610\|2 | ···················· | ···················· | ···················· | ···················· | NNNNNNNNNNNNNNNNNNNN | NNNNNNNNNNNNNNNNNNNN | 1 |
| M | 9275 | hCoV-19/Wales/PHWC-16DE60/2020\|EPI_ISL_585631\|2020 | ···················· | ···················· | ···················· | ···················· | NNNNNNNNNNNNNNNNNNNN | ···················· | 1 |
| M | 9278 | hCoV-19/Wales/PHWC-16DE9D/2020\|EPI_ISL_585634\|2020 | ···················· | ···················· | ···················· | ···················· | NNNNNNNNNNNNNNNNNNNN | ······NNNNNNNNN····· | 1 |
| M | 9281 | hCoV-19/Wales/PHWC-16DECA/2020\|EPI_ISL_585637\|2020 | ···················· | ···················· | ···················· | ···················· | NNNN················ | ···················· | 1 |
| M | 9282 | hCoV-19/Wales/PHWC-16DED9/2020\|EPI_ISL_585638\|2020 | ···················· | ···················· | ···················· | ···················· | NNNNNNNNNNNNNNNNNNNN | NNNNNNNNNNNNNNNNNNNN | 1 |
| M | 9283 | hCoV-19/Wales/PHWC-16DEF7/2020\|EPI_ISL_585639\|2020 | ···················· | ···················· | ···················· | ···················· | NNNN················ | ···················· | 1 |
| M | 9284 | hCoV-19/Wales/PHWC-16DF03/2020\|EPI_ISL_585640\|2020 | ···················· | ···················· | ···················· | ···················· | NNNNNNNNNNNNNNNNNNNN | ···················· | 1 |
| M | 9286 | hCoV-19/Wales/PHWC-16DF8B/2020\|EPI_ISL_585646\|2020 | ···················· | ···················· | ···················· | ···················· | NNNNNNNNNNNNNNNNNNNN | NNNNNNNNNNNNNNNNNNNN | 1 |
| M | 9289 | hCoV-19/Wales/PHWC-16DFD6/2020\|EPI_ISL_585651\|2020 | ···················· | ···················· | ···················· | ···················· | NNNNNNNNNNNNNNNNNNNN | NNNNNNNNNNNNNNNNNNNN | 1 |
| M | 9290 | hCoV-19/Wales/PHWC-16E166/2020\|EPI_ISL_585675\|2020 | ···················· | ···················· | ···················· | ···················· | NNNNNNNNNNNNNNNNNNNN | NNNNNNNNNNNNNNNNNNNN | 1 |
| M | 9291 | hCoV-19/Wales/PHWC-16E175/2020\|EPI_ISL_585676\|2020 | ···················· | ···················· | ···················· | ···················· | NNNNNNNNNNNNNNNNNNNN | ···················· | 1 |
| M | 9294 | hCoV-19/Wales/PHWC-16E1A2/2020\|EPI_ISL_585679\|2020 | ···················· | ···················· | ···················· | ···················· | NNNNNNNNNNNNNNNNNNNN | NNNNNNNNNNNNNNNNNNNN | 1 |
| M | 9295 | hCoV-19/Wales/PHWC-16E1B1/2020\|EPI_ISL_585680\|2020 | ···················· | ···················· | ···················· | ···················· | NNNNNNNNNNNNNNNNNNNN | ···················· | 1 |
| M | 9296 | hCoV-19/Wales/PHWC-16E1C0/2020\|EPI_ISL_585681\|2020 | ···················· | ···················· | ···················· | ···················· | NNNNNNNNNNNNNNNNNNNN | NNNNNNNNNNNNNNNNNNNN | 1 |
| M | 9372 | hCoV-19/Australia/VIC14888/2020\|EPI_ISL_592819\|202 | ···················· | ···················· | ···················· | ···················· | NNNNNNNNNNNNNNNNNNNN | ···················· | 1 |
| M | 9502 | hCoV-19/Wales/PHWC-16E01E/2020\|EPI_ISL_585655\|2020 | ···················· | ···················· | ···················· | ···················· | NNNNNNNNNNNNNNNNNNNN | NNNNNNNNNNNNNNNNNNNN | 1 |
| M | 9504 | hCoV-19/Wales/PHWC-16E04B/2020\|EPI_ISL_585658\|2020 | ···················· | ···················· | ···················· | ···················· | NNNNNNNNNNNNNNNNNNNN | NNNNNNNNNNNNNNNNNNNN | 1 |
| M | 9506 | hCoV-19/Wales/PHWC-16E069/2020\|EPI_ISL_585660\|2020 | ···················· | ···················· | ···················· | ···················· | NNNNNNNNNNNNNNNNNNNN | ···················· | 1 |
| M | 9507 | hCoV-19/Wales/PHWC-16E078/2020\|EPI_ISL_585661\|2020 | ···················· | ···················· | ···················· | ···················· | NNNNNNNNNNNNNNNNNNNN | ···················· | 1 |
| M | 9509 | hCoV-19/Wales/PHWC-16E096/2020\|EPI_ISL_585663\|2020 | ···················· | ···················· | ···················· | ···················· | NNNNNNNNNNNNNNNNNNNN | ···················· | 1 |
| M | 9510 | hCoV-19/Wales/PHWC-16E0A5/2020\|EPI_ISL_585664\|2020 | ···················· | ····A··············· | ···················· | ···················· | NNNNNNNNNNNNNNNNNNNN | ···················· | 1 |
| M | 9512 | hCoV-19/Wales/PHWC-16E0C3/2020\|EPI_ISL_585666\|2020 | ···················· | ···················· | ···················· | ···················· | NNNNNNNNNNNNNNNNNNNN | NNNNNNNNNNNNNNNNNNNN | 1 |
| M | 9514 | hCoV-19/Wales/PHWC-16E0E1/2020\|EPI_ISL_585668\|2020 | ···················· | ···················· | ···················· | ···················· | NNNNNNNNNNNNNNNNNNNN | NNNNNNNNNNNNNNNNNNNN | 1 |
| M | 9517 | hCoV-19/Wales/PHWC-16E12A/2020\|EPI_ISL_585672\|2020 | ···················· | ···················· | ···················· | ···················· | NNNNNNNNNNNNNNNNNNNN | ···················· | 1 |
| M | 9522 | hCoV-19/Wales/PHWC-16E236/2020\|EPI_ISL_585687\|2020 | ···················· | ···················· | ···················· | ···················· | NNNNNNNNNNNNNNNNNNNN | NNNNNNNNNNNNNNNNNNNN | 1 |
| N | 79 | hCoV-19/Australia/VIC14120/2020\|EPI_ISL_603048\|202 | ···················· | ···················· | ···················· | ···················· | NNNNNNNNNNNNNNNNNNNN | ···················· | 1 |
| N | 158 | hCoV-19/USA/WI-UW-1727/2020\|EPI_ISL_605600\|2020-09 | ···················· | ···················· | ···················· | ···················· | NNNNNNNNNNNNNNNNNNNN | NNNNNNNNNNNNNNNNNNNN | 1 |
| N | 159 | hCoV-19/USA/WI-UW-1728/2020\|EPI_ISL_605601\|2020-09 | ···················· | ···················· | ···················· | ···················· | NNNNNNNNNNNNNNNNNNNN | NNNNNNNNNNNNNNNNNNNN | 1 |
| N | 160 | hCoV-19/USA/WI-UW-1729/2020\|EPI_ISL_605602\|2020-09 | ···················· | ···················· | ···················· | ···················· | NNNNNNNNNNNNNNNNNNNN | NNNNNNNNNNNNNNNNNNNN | 1 |
| N | 161 | hCoV-19/USA/WI-UW-1730/2020\|EPI_ISL_605603\|2020-09 | ···················· | ···················· | ···················· | ···················· | NNNNNNNNNNNNNNNNNNNN | NNNNNNNNNNNNNNNNNNNN | 1 |

| | | | | | | | | |
|---|---|---|---|---|---|---|---|---|
| N | 162 hCoV-19/USA/WI-UW-1731/2020\|EPI_ISL_605604\|2020-09 | ···················· | ···················· | ···················· | ···················· | NNNNNNNNNNNNNNNNNNNN | NNNNNNNNNNNNNNNNNNNN | 1 |
| N | 163 hCoV-19/USA/WI-UW-1732/2020\|EPI_ISL_605605\|2020-09 | ···················· | ···················· | ···················· | ···················· | NNNNNNNNNNNNNNNNNNNN | NNNNNNNNNNNNNNNNNNNN | 1 |
| N | 164 hCoV-19/USA/WI-UW-1733/2020\|EPI_ISL_605606\|2020-09 | ···················· | ···················· | ···················· | ···················· | NNNNNNNNNNNNNNNNNNNN | NNNNNNNNNNNNNNNNNNNN | 1 |
| N | 165 hCoV-19/USA/WI-UW-1734/2020\|EPI_ISL_605607\|2020-09 | ···················· | ···················· | ···················· | ···················· | NNNNNNNNNNNNNNNNNNNN | NNNNNNNNNNNNNNNNNNNN | 1 |
| N | 166 hCoV-19/USA/WI-UW-1735/2020\|EPI_ISL_605608\|2020-09 | ···················· | ···················· | ···················· | ···················· | NNNNNNNNNNNNNNNNNNNN | NNNNNNNNNNNNNNNNNNNN | 1 |
| N | 167 hCoV-19/USA/WI-UW-1736/2020\|EPI_ISL_605609\|2020-09 | ···················· | ···················· | ···················· | ···················· | NNNNNNNNNNNNNNNNNNNN | NNNNNNNNNNNNNNNNNNNN | 1 |
| N | 168 hCoV-19/USA/WI-UW-1737/2020\|EPI_ISL_605610\|2020-09 | ···················· | ···················· | ···················· | ···················· | NNNNNNNNNNNNNNNNNNNN | NNNNNNNNNNNNNNNNNNNN | 1 |
| N | 169 hCoV-19/USA/WI-UW-1738/2020\|EPI_ISL_605611\|2020-09 | ···················· | ···················· | ···················· | ···················· | NNNNNNNNNNNNNNNNNNNN | NNNNNNNNNNNNNNNNNNNN | 1 |
| N | 170 hCoV-19/USA/WI-UW-1739/2020\|EPI_ISL_605612\|2020-09 | ···················· | ···················· | ···················· | ···················· | NNNNNNNNNNNNNNNNNNNN | NNNNNNNNNNNNNNNNNNNN | 1 |
| N | 177 hCoV-19/USA/WI-UW-1746/2020\|EPI_ISL_605619\|2020-09 | ···················· | ···················· | ···················· | ···················· | NNNNNNNNNNNNNNNNNNNN | NNNNNNNNNNNNNNNNNNNN | 1 |
| N | 196 hCoV-19/USA/WI-UW-1766/2020\|EPI_ISL_605639\|2020-09 | ···················· | ·············A······ | ···················· | ···················· | NNNNNNNNNNNNNNNNNNNN | NNNNNNNNNNNNNNNNNNNN | 1 |
| N | 200 hCoV-19/USA/WI-UW-1772/2020\|EPI_ISL_605644\|2020-09 | ···················· | ···················· | ···················· | ···················· | NNNNNNNNNNNNNNNNNNNN | NNNNNNNNNNNNNNNNNNNN | 1 |
| N | 201 hCoV-19/USA/WI-UW-1773/2020\|EPI_ISL_605645\|2020-09 | ···················· | ···················· | ···················· | ···················· | NNNNNNNNNNNNNNNNNNNN | NNNNNNNNNNNNNNNNNNNN | 1 |
| N | 206 hCoV-19/USA/WI-UW-1778/2020\|EPI_ISL_605650\|2020-09 | ···················· | ···················· | ···················· | ···················· | NNNNNNNNNNNNNNNNNNNN | NNNNNNNNNNNNNNNNNNNN | 1 |
| N | 212 hCoV-19/USA/WI-UW-1784/2020\|EPI_ISL_605656\|2020-09 | ···················· | ···················· | ···················· | ···················· | NNNNNNNNNNNNNNNNNNNN | NNNNNNNNNNNNNNNNNNNN | 1 |
| N | 213 hCoV-19/USA/WI-UW-1785/2020\|EPI_ISL_605657\|2020-09 | ···················· | ···················· | ···················· | ···················· | NNNNNNNNNNNNNNNNNNNN | NNNNNNNNNNNNNNNNNNNN | 1 |
| N | 214 hCoV-19/USA/WI-UW-1787/2020\|EPI_ISL_605659\|2020-09 | ···················· | ···················· | ···················· | ···················· | NNNNNNNNNNNNNNNNNNNN | NNNNNNNNNNNNNNNNNNNN | 1 |
| N | 215 hCoV-19/USA/WI-UW-1793/2020\|EPI_ISL_605665\|2020-09 | ···················· | ···················· | ···················· | ···················· | NNNNNNNNNNNNNNNNNNNN | NNNNNNNNNNNNNNNNNNNN | 1 |
| N | 216 hCoV-19/USA/WI-UW-1794/2020\|EPI_ISL_605666\|2020-09 | ···················· | ···················· | ···················· | ···················· | NNNNNNNNNNNNNNNNNNNN | NNNNNNNNNNNNNNNNNNNN | 1 |
| N | 217 hCoV-19/USA/WI-UW-1795/2020\|EPI_ISL_605667\|2020-09 | ···················· | ···················· | ···················· | ···················· | NNNNNNNNNNNNNNNNNNNN | NNNNNNNNNNNNNNNNNNNN | 1 |
| N | 218 hCoV-19/USA/WI-UW-1796/2020\|EPI_ISL_605668\|2020-09 | ···················· | ···················· | ···················· | ···················· | NNNNNNNNNNNNNNNNNNNN | NNNNNNNNNNNNNNNNNNNN | 1 |
| N | 219 hCoV-19/USA/WI-UW-1797/2020\|EPI_ISL_605669\|2020-09 | ···················· | ···················· | ···················· | ···················· | NNNNNNNNNNNNNNNNNNNN | NNNNNNNNNNNNNNNNNNNN | 1 |
| N | 221 hCoV-19/USA/WI-UW-1819/2020\|EPI_ISL_605691\|2020-09 | ···················· | ···················· | ···················· | ···················· | NNNNNNNNNNNNNNNNNNNN | NNNNNNNNNNNNNNNNNNNN | 1 |
| N | 222 hCoV-19/USA/WI-UW-1821/2020\|EPI_ISL_605693\|2020-09 | ···················· | ···················· | ···················· | ···················· | NNNNNNNNNNNNNNNNNNNN | NNNNNNNNNNNNNNNNNNNN | 1 |
| N | 223 hCoV-19/USA/WI-UW-1823/2020\|EPI_ISL_605695\|2020-09 | ···················· | ···················· | ···················· | ···················· | NNNNNNNNNNNNNNNNNNNN | NNNNNNNNNNNNNNNNNNNN | 1 |
| N | 224 hCoV-19/USA/WI-UW-1824/2020\|EPI_ISL_605696\|2020-09 | ···················· | ···················· | ···················· | ···················· | NNNNNNNNNNNNNNNNNNNN | NNNNNNNNNNNNNNNNNNNN | 1 |
| N | 225 hCoV-19/USA/WI-UW-1825/2020\|EPI_ISL_605697\|2020-09 | ···················· | ···················· | ···················· | ···················· | NNNNNNNNNNNNNNNNNNNN | NNNNNNNNNNNNNNNNNNNN | 1 |
| N | 226 hCoV-19/USA/WI-UW-1826/2020\|EPI_ISL_605698\|2020-09 | ···················· | ···················· | ···················· | ···················· | NNNNNNNNNNNNNNNNNNNN | NNNNNNNNNNNNNNNNNNNN | 1 |
| N | 256 hCoV-19/Hong Kong/HKPU-5116/2020\|EPI_ISL_610199\|20 | ···················· | ···················· | ···················· | ···················· | NNNNNNNNNNNNNNNNNNNN | NNNNNNNNNNNNNNNNNNNN | 1 |
| N | 489 hCoV-19/USA/WI-UW-1788/2020\|EPI_ISL_605660\|2020-09 | ···················· | ···················· | ···················· | ···················· | NNNNNNNNNNNNNNNNNNNN | NNNNNNNNNNNNNNNNNNNN | 1 |
| N | 490 hCoV-19/USA/WI-UW-1789/2020\|EPI_ISL_605661\|2020-09 | ···················· | ···················· | ···················· | ···················· | NNNNNNNNNNNNNNNNNNNN | NNNNNNNNNNNNNNNNNNNN | 1 |
| N | 491 hCoV-19/USA/WI-UW-1790/2020\|EPI_ISL_605662\|2020-09 | ···················· | ···················· | ···················· | ···················· | NNNNNNNNNNNNNNNNNNNN | NNNNNNNNNNNNNNNNNNNN | 1 |
| N | 492 hCoV-19/USA/WI-UW-1791/2020\|EPI_ISL_605663\|2020-09 | ···················· | ···················· | ···················· | ···················· | NNNNNNNNNNNNNNNNNNNN | NNNNNNNNNNNNNNNNNNNN | 1 |
| N | 493 hCoV-19/USA/WI-UW-1792/2020\|EPI_ISL_605664\|2020-09 | ···················· | ···················· | ···················· | ···················· | NNNNNNNNNNNNNNNNNNNN | NNNNNNNNNNNNNNNNNNNN | 1 |
| N | 494 hCoV-19/USA/WI-UW-1798/2020\|EPI_ISL_605670\|2020-09 | ···················· | ···················· | ···················· | ···················· | NNNNNNNNNNNNNNNNNNNN | NNNNNNNNNNNNNNNNNNNN | 1 |
| N | 495 hCoV-19/USA/WI-UW-1799/2020\|EPI_ISL_605671\|2020-09 | ···················· | ···················· | ···················· | ···················· | NNNNNNNNNNNNNNNNNNNN | NNNNNNNNNNNNNNNNNNNN | 1 |
| N | 496 hCoV-19/USA/WI-UW-1800/2020\|EPI_ISL_605672\|2020-09 | ···················· | ···················· | ···················· | ···················· | NNNNNNNNNNNNNNNNNNNN | NNNNNNNNNNNNNNNNNNNN | 1 |
| N | 497 hCoV-19/USA/WI-UW-1801/2020\|EPI_ISL_605673\|2020-09 | ···················· | ···················· | ···················· | ···················· | NNNNNNNNNNNNNNNNNNNN | NNNNNNNNNNNNNNNNNNNN | 1 |
| N | 498 hCoV-19/USA/WI-UW-1802/2020\|EPI_ISL_605674\|2020-09 | ···················· | ···················· | ···················· | ···················· | NNNNNNNNNNNNNNNNNNNN | NNNNNNNNNNNNNNNNNNNN | 1 |
| N | 499 hCoV-19/USA/WI-UW-1803/2020\|EPI_ISL_605675\|2020-09 | ···················· | ···················· | ···················· | ···················· | NNNNNNNNNNNNNNNNNNNN | NNNNNNNNNNNNNNNNNNNN | 1 |
| N | 502 hCoV-19/USA/WI-UW-1806/2020\|EPI_ISL_605678\|2020-09 | ···················· | ···················· | ···················· | ···················· | NNNNNNNNNNNNNNNNNNNN | NNNNNNNNNNNNNNNNNNNN | 1 |
| N | 503 hCoV-19/USA/WI-UW-1807/2020\|EPI_ISL_605679\|2020-09 | ···················· | ···················· | ···················· | ···················· | NNNNNNNNNNNNNNNNNNNN | NNNNNNNNNNNNNNNNNNNN | 1 |
| N | 504 hCoV-19/USA/WI-UW-1808/2020\|EPI_ISL_605680\|2020-09 | ···················· | ···················· | ···················· | ···················· | NNNNNNNNNNNNNNNNNNNN | NNNNNNNNNNNNNNNNNNNN | 1 |
| N | 506 hCoV-19/USA/WI-UW-1810/2020\|EPI_ISL_605682\|2020-09 | ···················· | ···················· | ···················· | ···················· | NNNNNNNNNNNNNNNNNNNN | NNNNNNNNNNNNNNNNNNNN | 1 |
| N | 507 hCoV-19/USA/WI-UW-1811/2020\|EPI_ISL_605683\|2020-09 | ···················· | ···················· | ···················· | ···················· | NNNNNNNNNNNNNNNNNNNN | NNNNNNNNNNNNNNNNNNNN | 1 |
| N | 508 hCoV-19/USA/WI-UW-1812/2020\|EPI_ISL_605684\|2020-09 | ···················· | ···················· | ···················· | ···················· | NNNNNNNNNNNNNNNNNNNN | NNNNNNNNNNNNNNNNNNNN | 1 |
| N | 509 hCoV-19/USA/WI-UW-1813/2020\|EPI_ISL_605685\|2020-09 | ···················· | ···················· | ···················· | ···················· | NNNNNNNNNNNNNNNNNNNN | NNNNNNNNNNNNNNNNNNNN | 1 |
| N | 510 hCoV-19/USA/WI-UW-1815/2020\|EPI_ISL_605687\|2020-09 | ···················· | ···················· | ···················· | ···················· | NNNNNNNNNNNNNNNNNNNN | NNNNNNNNNNNNNNNNNNNN | 1 |
| N | 511 hCoV-19/USA/WI-UW-1816/2020\|EPI_ISL_605688\|2020-09 | ···················· | ···················· | ···················· | ···················· | NNNNNNNNNNNNNNNNNNNN | NNNNNNNNNNNNNNNNNNNN | 1 |
| N | 512 hCoV-19/USA/WI-UW-1817/2020\|EPI_ISL_605689\|2020-09 | ···················· | ···················· | ···················· | ···················· | NNNNNNNNNNNNNNNNNNNN | NNNNNNNNNNNNNNNNNNNN | 1 |
| N | 702 hCoV-19/USA/WI-UW-1637/2020\|EPI_ISL_605511\|2020-09 | ···················· | ···················· | ···················· | ···················· | NNNNNNNNNNNNNNNNNNNN | NNNNNNNNNNNNNNNNNNNN | 1 |
| N | 706 hCoV-19/USA/WI-UW-1641/2020\|EPI_ISL_605515\|2020-09 | ···················· | ···················· | ···················· | ···················· | NNNNNNNNNNNNNNNNNNNN | NNNNNNNNNNNNNNNNNNNN | 1 |
| N | 708 hCoV-19/USA/WI-UW-1643/2020\|EPI_ISL_605517\|2020-09 | ···················· | ···················· | ···················· | ···················· | NNNNNNNNNNNNNNNNNNNN | NNNNNNNNNNNNNNNNNNNN | 1 |
| N | 709 hCoV-19/USA/WI-UW-1644/2020\|EPI_ISL_605518\|2020-09 | ···················· | ···················· | ···················· | ···················· | NNNNNNNNNNNNNNNNNNNN | NNNNNNNNNNNNNNNNNNNN | 1 |
| N | 711 hCoV-19/USA/WI-UW-1646/2020\|EPI_ISL_605520\|2020-09 | ···················· | ···················· | ···················· | ···················· | NNNNNNNNNNNNNNNNNNNN | NNNNNNNNNNNNNNNNNNNN | 1 |
| N | 720 hCoV-19/USA/WI-UW-1655/2020\|EPI_ISL_605529\|2020-09 | ···················· | ···················· | ···················· | ···················· | NNNNNNNNNNNNNNNNNNNN | NNNNNNNNNNNNNNNNNNNN | 1 |
| N | 726 hCoV-19/USA/WI-UW-1661/2020\|EPI_ISL_605535\|2020-09 | ···················· | ···················· | ···················· | ···················· | NNNNNNNNNNNNNNNNNNNN | NNNNNNNNNNNNNNNNNNNN | 1 |
| N | 727 hCoV-19/USA/WI-UW-1662/2020\|EPI_ISL_605536\|2020-09 | ···················· | ···················· | ···················· | ···················· | NNNNNNNNNNNNNNNNNNNN | NNNNNNNNNNNNNNNNNNNN | 1 |
| N | 729 hCoV-19/USA/WI-UW-1664/2020\|EPI_ISL_605538\|2020-09 | ···················· | ···················· | ···················· | ···················· | NNNNNNNNNNNNNNNNNNNN | NNNNNNNNNNNNNNNNNNNN | 1 |
| N | 733 hCoV-19/USA/WI-UW-1668/2020\|EPI_ISL_605542\|2020-09 | ···················· | ···················· | ···················· | ···················· | NNNNNNNNNNNNNNNNNNNN | NNNNNNNNNNNNNNNNNNNN | 1 |
| N | 735 hCoV-19/USA/WI-UW-1670/2020\|EPI_ISL_605544\|2020-09 | ···················· | ···················· | ···················· | ···················· | NNNNNNNNNNNNNNNNNNNN | NNNNNNNNNNNNNNNNNNNN | 1 |
| N | 738 hCoV-19/USA/WI-UW-1673/2020\|EPI_ISL_605547\|2020-09 | ···················· | ···················· | ···················· | ···················· | NNNNNNNNNNNNNNNNNNNN | NNNNNNNNNNNNNNNNNNNN | 1 |
| N | 741 hCoV-19/USA/WI-UW-1697/2020\|EPI_ISL_605570\|2020-09 | ···················· | ···················· | ···················· | ···················· | NNNNNNNNNNNNNNNNNNNN | NNNNNNNNNNNNNNNNNNNN | 1 |
| N | 742 hCoV-19/USA/WI-UW-1698/2020\|EPI_ISL_605571\|2020-09 | ···················· | ···················· | ···················· | ···················· | NNNNNNNNNNNNNNNNNNNN | NNNNNNNNNNNNNNNNNNNN | 1 |
| N | 744 hCoV-19/USA/WI-UW-1700/2020\|EPI_ISL_605573\|2020-09 | ···················· | ···················· | ···················· | ···················· | NNNNNNNNNNNNNNNNNNNN | NNNNNNNNNNNNNNNNNNNN | 1 |
| N | 745 hCoV-19/USA/WI-UW-1701/2020\|EPI_ISL_605574\|2020-09 | ···················· | ···················· | ···················· | ···················· | NNNNNNNNNNNNNNNNNNNN | NNNNNNNNNNNNNNNNNNNN | 1 |
| N | 747 hCoV-19/USA/WI-UW-1703/2020\|EPI_ISL_605576\|2020-09 | ···················· | ·············A······ | ···················· | ···················· | NNNNNNNNNNNNNNNNNNNN | NNNNNNNNNNNNNNNNNNNN | 1 |
| N | 750 hCoV-19/USA/WI-UW-1706/2020\|EPI_ISL_605579\|2020-09 | ···················· | ···················· | ···················· | ···················· | NNNNNNNNNNNNNNNNNNNN | NNNNNNNNNNNNNNNNNNNN | 1 |
| N | 752 hCoV-19/USA/WI-UW-1708/2020\|EPI_ISL_605581\|2020-09 | ···················· | ···················· | ···················· | ···················· | NNNNNNNNNNNNNNNNNNNN | NNNNNNNNNNNNNNNNNNNN | 1 |
| N | 755 hCoV-19/USA/WI-UW-1711/2020\|EPI_ISL_605584\|2020-09 | ···················· | ···················· | ···················· | ···················· | NNNNNNNNNNNNNNNNNNNN | ···················· | 1 |
| N | 756 hCoV-19/USA/WI-UW-1713/2020\|EPI_ISL_605586\|2020-09 | ···················· | ···················· | ···················· | ···················· | NNNNNNNNNNNNNNNNNNNN | NNNNNNNNNNNNNNNNNNNN | 1 |
| N | 759 hCoV-19/USA/WI-UW-1716/2020\|EPI_ISL_605589\|2020-09 | ···················· | ···················· | ···················· | ···················· | NNNNNNNNNNNNNNNNNNNN | NNNNNNNNNNNNNNNNNNNN | 1 |
| N | 760 hCoV-19/USA/WI-UW-1717/2020\|EPI_ISL_605590\|2020-09 | ···················· | ···················· | ···················· | ···················· | NNNNNNNNNNNNNNNNNNNN | NNNNNNNNNNNNNNNNNNNN | 1 |
| N | 761 hCoV-19/USA/WI-UW-1718/2020\|EPI_ISL_605591\|2020-09 | ···················· | ·············A······ | ···················· | ···················· | NNNNNNNNNNNNNNNNNNNN | NNNNNNNNNNNNNNNNNNNN | 1 |
| N | 762 hCoV-19/USA/WI-UW-1719/2020\|EPI_ISL_605592\|2020-09 | ···················· | ···················· | ···················· | ···················· | NNNNNNNNNNNNNNNNNNNN | NNNNNNNNNNNNNNNNNNNN | 1 |
| N | 763 hCoV-19/USA/WI-UW-1720/2020\|EPI_ISL_605593\|2020-09 | ···················· | ···················· | ···················· | ···················· | NNNNNNNNNNNNNNNNNNNN | NNNNNNNNNNNNNNNNNNNN | 1 |
| N | 764 hCoV-19/USA/WI-UW-1721/2020\|EPI_ISL_605594\|2020-09 | ···················· | ···················· | ···················· | ···················· | NNNNNNNNNNNNNNNNNNNN | NNNNNNNNNNNNNNNNNNNN | 1 |
| N | 765 hCoV-19/USA/WI-UW-1722/2020\|EPI_ISL_605595\|2020-09 | ···················· | ···················· | ···················· | ···················· | NNNNNNNNNNNNNNNNNNNN | NNNNNNNNNNNNNNNNNNNN | 1 |
| N | 766 hCoV-19/USA/WI-UW-1723/2020\|EPI_ISL_605596\|2020-09 | ···················· | ···················· | ···················· | ···················· | NNNNNNNNNNNNNNNNNNNN | NNNNNNNNNNNNNNNNNNNN | 1 |
| N | 768 hCoV-19/USA/WI-UW-1726/2020\|EPI_ISL_605599\|2020-09 | ···················· | ···················· | ···················· | ···················· | NNNNNNNNNNNNNNNNNNNN | NNNNNNNNNNNNNNNNNNNN | 1 |

| | | | | | | | | |
|---|---|---|---|---|---|---|---|---|
| N | 772 hCoV-19/England/CAMC-A57D72/2020\|EPI_ISL_596987\|20 | ···················· | ···················· | ···················· | ···················· | NNNNNNNNNNNNNNNNNNNN | NNNNNNNNNNNNNNNNNNNN | 1 |
| N | 1003 hCoV-19/USA/IL-UW-1617/2020\|EPI_ISL_605495\|2020-09 | ···················· | ···················· | ···················· | ···················· | NNNNNNNNNNNNNNNNNNNN | NNNNNNNNNNNNNNNNNNNN | 1 |
| N | 1006 hCoV-19/USA/WI-UW-1676/2020\|EPI_ISL_605550\|2020-09 | ···················· | ···················· | ···················· | ···················· | NNNNNNNNNNNNNNNNNNNN | NNNNNNNNNNNNNNNNNNNN | 1 |
| N | 1009 hCoV-19/USA/WI-UW-1681/2020\|EPI_ISL_605554\|2020-09 | ···················· | ···················· | ···················· | ···················· | NNNNNNNNNNNNNNNNNNNN | NNNNNNNNNNNNNNNNNNNN | 1 |
| N | 1010 hCoV-19/USA/WI-UW-1682/2020\|EPI_ISL_605555\|2020-09 | ···················· | ···················· | ···················· | ···················· | NNNNNNNNNNNNNNNNNNNN | NNNNNNNNNNNNNNNNNNNN | 1 |
| N | 1011 hCoV-19/USA/WI-UW-1683/2020\|EPI_ISL_605556\|2020-09 | ···················· | ·············A······ | ···················· | ···················· | NNNNNNNNNNNNNNNNNNNN | NNNNNNNNNNNNNNNNNNNN | 1 |
| N | 1012 hCoV-19/USA/WI-UW-1684/2020\|EPI_ISL_605557\|2020-09 | ···················· | ···················· | ···················· | ···················· | NNNNNNNNNNNNNNNNNNNN | NNNNNNNNNNNNNNNNNNNN | 1 |
| N | 1014 hCoV-19/USA/WI-UW-1686/2020\|EPI_ISL_605559\|2020-09 | ···················· | ···················· | ···················· | ···················· | NNNNNNNNNNNNNNNNNNNN | NNNNNNNNNNNNNNNNNNNN | 1 |
| N | 1015 hCoV-19/USA/WI-UW-1687/2020\|EPI_ISL_605560\|2020-09 | ···················· | ···················· | ···················· | ···················· | NNNNNNNNNNNNNNNNNNNN | NNNNNNNNNNNNNNNNNNNN | 1 |
| N | 1017 hCoV-19/USA/WI-UW-1689/2020\|EPI_ISL_605562\|2020-09 | ···················· | ···················· | ···················· | ···················· | NNNNNNNNNNNNNNNNNNNN | NNNNNNNNNNNNNNNNNNNN | 1 |
| N | 1019 hCoV-19/USA/WI-UW-1691/2020\|EPI_ISL_605564\|2020-09 | ···················· | ···················· | ···················· | ···················· | NNNNNNNNNNNNNNNNNNNN | NNNNNNNNNNNNNNNNNNNN | 1 |
| N | 1020 hCoV-19/USA/WI-UW-1692/2020\|EPI_ISL_605565\|2020-09 | ···················· | ···················· | ···················· | ···················· | NNNNNNNNNNNNNNNNNNNN | NNNNNNNNNNNNNNNNNNNN | 1 |
| N | 1023 hCoV-19/USA/WI-UW-1695/2020\|EPI_ISL_605568\|2020-09 | ···················· | ···················· | ···················· | ···················· | NNNNNNNNNNNNNNNNNNNN | NNNNNNNNNNNNNNNNNNNN | 1 |
| N | 1239 hCoV-19/USA/WI-UW-1529/2020\|EPI_ISL_605407\|2020-09 | ···················· | ···················· | ···················· | ···················· | NNNNNNNNNNNNNNNNNNNN | NNNNNNNNNNNNNNNNNNNN | 1 |
| N | 1245 hCoV-19/USA/WI-UW-1535/2020\|EPI_ISL_605413\|2020-09 | ···················· | ···················· | ···················· | ···················· | NNNNNNNNNNNNNNNNNNNN | NNNNNNNNNNNNNNNNNNNN | 1 |
| N | 1246 hCoV-19/USA/WI-UW-1536/2020\|EPI_ISL_605414\|2020-09 | ···················· | ···················· | ···················· | ···················· | NNNNNNNNNNNNNNNNNNNN | NNNNNNNNNNNNNNNNNNNN | 1 |
| N | 1251 hCoV-19/USA/WI-UW-1572/2020\|EPI_ISL_605450\|2020-09 | ···················· | ···················· | ···················· | ···················· | NNNNNNNNNNNNNNNNNNNN | NNNNNNNNNNNNNNNNNNNN | 1 |
| N | 1252 hCoV-19/USA/WI-UW-1578/2020\|EPI_ISL_605455\|2020-09 | ···················· | ···················· | ···················· | ···················· | NNNNNNNNNNNNNNNNNNNN | NNNNNNNNNNNNNNNNNNNN | 1 |
| N | 1253 hCoV-19/USA/WI-UW-1579/2020\|EPI_ISL_605456\|2020-09 | ···················· | ···················· | ···················· | ···················· | NNNNNNNNNNNNNNNNNNNN | NNNNNNNNNNNNNNNNNNNN | 1 |
| N | 1254 hCoV-19/USA/WI-UW-1580/2020\|EPI_ISL_605457\|2020-09 | ···················· | ···················· | ···················· | ···················· | NNNNNNNNNNNNNNNNNNNN | NNNNNNNNNNNNNNNNNNNN | 1 |
| N | 1255 hCoV-19/USA/WI-UW-1581/2020\|EPI_ISL_605458\|2020-09 | ···················· | ···················· | ···················· | ···················· | NNNNNNNNNNNNNNNNNNNN | NNNNNNNNNNNNNNNNNNNN | 1 |
| N | 1256 hCoV-19/USA/WI-UW-1582/2020\|EPI_ISL_605459\|2020-09 | ···················· | ···················· | ···················· | ···················· | NNNNNNNNNNNNNNNNNNNN | NNNNNNNNNNNNNNNNNNNN | 1 |
| N | 1257 hCoV-19/USA/WI-UW-1584/2020\|EPI_ISL_605461\|2020-09 | ···················· | ···················· | ···················· | ···················· | NNNNNNNNNNNNNNNNNNNN | NNNNNNNNNNNNNNNNNNNN | 1 |
| N | 1258 hCoV-19/USA/WI-UW-1585/2020\|EPI_ISL_605462\|2020-09 | ···················· | ···················· | ···················· | ···················· | NNNNNNNNNNNNNNNNNNNN | NNNNNNNNNNNNNNNNNNNN | 1 |
| N | 1259 hCoV-19/USA/WI-UW-1586/2020\|EPI_ISL_605463\|2020-09 | ···················· | ···················· | ···················· | ···················· | NNNNNNNNNNNNNNNNNNNN | NNNNNNNNNNNNNNNNNNNN | 1 |
| N | 1260 hCoV-19/USA/WI-UW-1587/2020\|EPI_ISL_605464\|2020-09 | ···················· | ···················· | ···················· | ···················· | NNNNNNNNNNNNNNNNNNNN | NNNNNNNNNNNNNNNNNNNN | 1 |
| N | 1265 hCoV-19/USA/WI-UW-1594/2020\|EPI_ISL_605471\|2020-09 | ···················· | ···················· | ···················· | ···················· | NNNNNNNNNNNNNNNNNNNN | NNNNNNNNNNNNNNNNNNNN | 1 |
| N | 1266 hCoV-19/USA/WI-UW-1596/2020\|EPI_ISL_605473\|2020-09 | ···················· | ···················· | ···················· | ···················· | NNNNNNNNNNNNNNNNNNNN | NNNNNNNNNNNNNNNNNNNN | 1 |
| N | 1271 hCoV-19/USA/WI-UW-1601/2020\|EPI_ISL_605478\|2020-09 | ···················· | ···················· | ···················· | ···················· | NNNNNNNNNNNNNNNNNNNN | NNNNNNNNNNNNNNNNNNNN | 1 |
| N | 1274 hCoV-19/USA/WI-UW-1604/2020\|EPI_ISL_605481\|2020-09 | ···················· | ···················· | ···················· | ···················· | NNNNNNNNNNNNNNNNNNNN | NNNNNNNNNNNNNNNNNNNN | 1 |
| N | 1518 hCoV-19/USA/WI-UW-1552/2020\|EPI_ISL_605430\|2020-09 | ···················· | ···················· | ···················· | ···················· | NNNNNNNNNNNNNNNNNNNN | NNNNNNNNNNNNNNNNNNNN | 1 |
| N | 1519 hCoV-19/USA/WI-UW-1553/2020\|EPI_ISL_605431\|2020-09 | ···················· | ···················· | ···················· | ···················· | NNNNNNNNNNNNNNNNNNNN | NNNNNNNNNNNNNNNNNNNN | 1 |
| N | 1520 hCoV-19/USA/WI-UW-1554/2020\|EPI_ISL_605432\|2020-09 | ···················· | ···················· | ···················· | ···················· | NNNNNNNNNNNNNNNNNNNN | NNNNNNNNNNNNNNNNNNNN | 1 |
| N | 1522 hCoV-19/USA/WI-UW-1556/2020\|EPI_ISL_605434\|2020-09 | ···················· | ···················· | ···················· | ···················· | NNNNNNNNNNNNNNNNNNNN | NNNNNNNNNNNNNNNNNNNN | 1 |
| N | 1523 hCoV-19/USA/WI-UW-1557/2020\|EPI_ISL_605435\|2020-09 | ···················· | ···················· | ···················· | ···················· | NNNNNNNNNNNNNNNNNNNN | NNNNNNNNNNNNNNNNNNNN | 1 |
| N | 1524 hCoV-19/USA/WI-UW-1558/2020\|EPI_ISL_605436\|2020-09 | ···················· | ···················· | ···················· | ···················· | NNNNNNNNNNNNNNNNNNNN | NNNNNNNNNNNNNNNNNNNN | 1 |
| N | 1525 hCoV-19/USA/WI-UW-1559/2020\|EPI_ISL_605437\|2020-09 | ···················· | ···················· | ···················· | ···················· | NNNNNNNNNNNNNNNNNNNN | NNNNNNNNNNNNNNNNNNNN | 1 |
| N | 1526 hCoV-19/USA/WI-UW-1560/2020\|EPI_ISL_605438\|2020-09 | ···················· | ···················· | ···················· | ···················· | NNNNNNNNNNNNNNNNNNNN | NNNNNNNNNNNNNNNNNNNN | 1 |
| N | 1528 hCoV-19/USA/WI-UW-1563/2020\|EPI_ISL_605441\|2020-09 | ···················· | ···················· | ···················· | ···················· | NNNNNNNNNNNNNNNNNNNN | NNNNNNNNNNNNNNNNNNNN | 1 |
| N | 1530 hCoV-19/USA/WI-UW-1565/2020\|EPI_ISL_605443\|2020-09 | ···················· | ···················· | ···················· | ···················· | NNNNNNNNNNNNNNNNNNNN | NNNNNNNNNNNNNNNNNNNN | 1 |
| N | 1531 hCoV-19/USA/WI-UW-1566/2020\|EPI_ISL_605444\|2020-09 | ···················· | ···················· | ···················· | ···················· | NNNNNNNNNNNNNNNNNNNN | NNNNNNNNNNNNNNNNNNNN | 1 |
| N | 1533 hCoV-19/USA/WI-UW-1568/2020\|EPI_ISL_605446\|2020-09 | ···················· | ···················· | ···················· | ···················· | NNNNNNNNNNNNNNNNNNNN | NNNNNNNNNNNNNNNNNNNN | 1 |
| N | 1534 hCoV-19/USA/WI-UW-1569/2020\|EPI_ISL_605447\|2020-09 | ···················· | ···················· | ···················· | ···················· | NNNNNNNNNNNNNNNNNNNN | NNNNNNNNNNNNNNNNNNNN | 1 |
| N | 1536 hCoV-19/USA/WI-UW-1571/2020\|EPI_ISL_605449\|2020-09 | ···················· | ···················· | ···················· | ···················· | NNNNNNNNNNNNNNNNNNNN | NNNNNNNNNNNNNNNNNNNN | 1 |
| N | 1549 hCoV-19/Australia/WA149/2020\|EPI_ISL_596712\|2020-0 | ···················· | ···················· | ···················· | ···················· | ···················· | ··········NNNNNN···· | 1 |
| N | 3519 hCoV-19/USA/WI-UW-1828/2020\|EPI_ISL_605700\|2020-09 | ···················· | ···················· | ···················· | ···················· | NNNNNNNNNNNNNNNNNNNN | NNNNNNNNNNNNNNNNNNNN | 1 |
| N | 3522 hCoV-19/USA/WI-UW-1831/2020\|EPI_ISL_605703\|2020-09 | ···················· | ···················· | ···················· | ···················· | NNNNNNNNNNNNNNNNNNNN | NNNNNNNNNNNNNNNNNNNN | 1 |
| N | 3528 hCoV-19/USA/WI-UW-1838/2020\|EPI_ISL_605710\|2020-09 | ···················· | ···················· | ···················· | ···················· | NNNNNNNNNNNNNNNNNNNN | NNNNNNNNNNNNNNNNNNNN | 1 |
| N | 3529 hCoV-19/USA/WI-UW-1839/2020\|EPI_ISL_605711\|2020-09 | ···················· | ···················· | ···················· | ···················· | NNNNNNNNNNNNNNNNNNNN | NNNNNNNNNNNNNNNNNNNN | 1 |
| N | 3538 hCoV-19/USA/WI-UW-1848/2020\|EPI_ISL_605720\|2020-09 | ···················· | ···················· | ···················· | ···················· | NNNNNNNNNNNNNNNNNNNN | NNNNNNNNNNNNNNNNNNNN | 1 |
| N | 3540 hCoV-19/USA/WI-UW-1850/2020\|EPI_ISL_605722\|2020-09 | ···················· | ···················· | ···················· | ···················· | NNNNNNNNNNNNNNNNNNNN | NNNNNNNNNNNNNNNNNNNN | 1 |
| N | 3541 hCoV-19/USA/WI-UW-1851/2020\|EPI_ISL_605723\|2020-09 | ···················· | ···················· | ···················· | ···················· | NNNNNNNNNNNNNNNNNNNN | NNNNNNNNNNNNNNNNNNNN | 1 |
| N | 3542 hCoV-19/USA/WI-UW-1852/2020\|EPI_ISL_605724\|2020-09 | ···················· | ···················· | ···················· | ···················· | NNNNNNNNNNNNNNNNNNNN | NNNNNNNNNNNNNNNNNNNN | 1 |
| N | 3543 hCoV-19/USA/WI-UW-1853/2020\|EPI_ISL_605725\|2020-09 | ···················· | ···················· | ···················· | ···················· | NNNNNNNNNNNNNNNNNNNN | NNNNNNNNNNNNNNNNNNNN | 1 |
| N | 3545 hCoV-19/USA/WI-UW-1855/2020\|EPI_ISL_605727\|2020-09 | ···················· | ···················· | ···················· | ···················· | NNNNNNNNNNNNNNNNNNNN | NNNNNNNNNNNNNNNNNNNN | 1 |
| N | 3546 hCoV-19/USA/WI-UW-1856/2020\|EPI_ISL_605728\|2020-09 | ···················· | ···················· | ···················· | ···················· | NNNNNNNNNNNNNNNNNNNN | NNNNNNNNNNNNNNNNNNNN | 1 |
| N | 3547 hCoV-19/USA/WI-UW-1857/2020\|EPI_ISL_605729\|2020-09 | ···················· | ···················· | ···················· | ···················· | NNNNNNNNNNNNNNNNNNNN | NNNNNNNNNNNNNNNNNNNN | 1 |
| N | 3548 hCoV-19/USA/WI-UW-1858/2020\|EPI_ISL_605730\|2020-09 | ···················· | ···················· | ···················· | ···················· | NNNNNNNNNNNNNNNNNNNN | NNNNNNNNNNNNNNNNNNNN | 1 |
| N | 3550 hCoV-19/USA/WI-UW-1860/2020\|EPI_ISL_605732\|2020-09 | ···················· | ···················· | ···················· | ···················· | NNNNNNNNNNNNNNNNNNNN | NNNNNNNNNNNNNNNNNNNN | 1 |
| N | 3551 hCoV-19/USA/WI-UW-1861/2020\|EPI_ISL_605733\|2020-09 | ···················· | ···················· | ···················· | ···················· | NNNNNNNNNNNNNNNNNNNN | NNNNNNNNNNNNNNNNNNNN | 1 |
| N | 3552 hCoV-19/USA/WI-UW-1862/2020\|EPI_ISL_605734\|2020-09 | ···················· | ···················· | ···················· | ···················· | NNNNNNNNNNNNNNNNNNNN | NNNNNNNNNNNNNNNNNNNN | 1 |
| N | 3555 hCoV-19/USA/WI-UW-1865/2020\|EPI_ISL_605737\|2020-09 | ···················· | ···················· | ···················· | ···················· | NNNNNNNNNNNNNNNNNNNN | NNNNNNNNNNNNNNNNNNNN | 1 |
| N | 3556 hCoV-19/USA/WI-UW-1866/2020\|EPI_ISL_605738\|2020-09 | ···················· | ···················· | ···················· | ···················· | NNNNNNNNNNNNNNNNNNNN | NNNNNNNNNNNNNNNNNNNN | 1 |
| N | 3557 hCoV-19/USA/WI-UW-1867/2020\|EPI_ISL_605739\|2020-09 | ···················· | ···················· | ···················· | ···················· | NNNNNNNNNNNNNNNNNNNN | NNNNNNNNNNNNNNNNNNNN | 1 |
| N | 3558 hCoV-19/USA/WI-UW-1868/2020\|EPI_ISL_605740\|2020-09 | ···················· | ···················· | ···················· | ···················· | NNNNNNNNNNNNNNNNNNNN | NNNNNNNNNNNNNNNNNNNN | 1 |
| N | 3559 hCoV-19/USA/WI-UW-1869/2020\|EPI_ISL_605741\|2020-09 | ···················· | ···················· | ···················· | ···················· | NNNNNNNNNNNNNNNNNNNN | NNNNNNNNNNNNNNNNNNNN | 1 |
| N | 3560 hCoV-19/USA/WI-UW-1870/2020\|EPI_ISL_605742\|2020-09 | ···················· | ···················· | ···················· | ···················· | NNNNNNNNNNNNNNNNNNNN | NNNNNNNNNNNNNNNNNNNN | 1 |
| N | 3561 hCoV-19/USA/WI-UW-1871/2020\|EPI_ISL_605743\|2020-09 | ···················· | ···················· | ···················· | ···················· | NNNNNNNNNNNNNNNNNNNN | NNNNNNNNNNNNNNNNNNNN | 1 |
| N | 3564 hCoV-19/USA/WI-UW-1875/2020\|EPI_ISL_605747\|2020-09 | ···················· | ···················· | ···················· | ···················· | NNNNNNNNNNNNNNNNNNNN | NNNNNNNNNNNNNNNNNNNN | 1 |
| N | 3567 hCoV-19/USA/WI-UW-1879/2020\|EPI_ISL_605750\|2020-09 | ···················· | ···················· | ···················· | ···················· | NNNNNNNNNNNNNNNNNNNN | NNNNNNNNNNNNNNNNNNNN | 1 |
| N | 3569 hCoV-19/USA/WI-UW-1881/2020\|EPI_ISL_605752\|2020-09 | ···················· | ···················· | ···················· | ···················· | NNNNNNNNNNNNNNNNNNNN | NNNNNNNNNNNNNNNNNNNN | 1 |
| N | 3570 hCoV-19/USA/WI-UW-1884/2020\|EPI_ISL_605754\|2020-09 | ···················· | ···················· | ···················· | ···················· | NNNNNNNNNNNNNNNNNNNN | NNNNNNNNNNNNNNNNNNNN | 1 |
| N | 3578 hCoV-19/USA/WI-UW-1893/2020\|EPI_ISL_605762\|2020-09 | ···················· | ···················· | ···················· | ···················· | NNNNNNNNNNNNNNNNNNNN | NNNNNNNNNNNNNNNNNNNN | 1 |
| N | 3659 hCoV-19/England/QEUH-A0C90F/2020\|EPI_ISL_599343\|20 | ···················· | NNNNNNNNNNNNNNNNNNNN | ···················· | ···················· | ···················· | ···················· | 1 |
| N | 3822 hCoV-19/USA/WI-UW-1906/2020\|EPI_ISL_605775\|2020-09 | ···················· | ···················· | ···················· | ···················· | NNNNNNNNNNNNNNNNNNNN | NNNNNNNNNNNNNNNNNNNN | 1 |
| N | 4083 hCoV-19/Hong Kong/HKPU-1166/2020\|EPI_ISL_610211\|20 | ···················· | ···················· | ···················· | ···················· | NNNNNNNNNNNNNNNNNNNN | NNNNNNNNNNNNNNNNNNNN | 1 |
| N | 4088 hCoV-19/Hong Kong/HKPU-4896/2020\|EPI_ISL_610217\|20 | ···················· | ···················· | ···················· | ···················· | NNNNNNNNNNNNNNNNNNNN | NNNNNNNNNNNNNNNNNNNN | 1 |
| N | 4089 hCoV-19/Hong Kong/HKPU-4954/2020\|EPI_ISL_610218\|20 | ···················· | ···················· | ···················· | ···················· | NNNNNNNNNNNNNNNNNNNN | NNNNNNNNNNNNNNNNNNNN | 1 |
| N | 4092 hCoV-19/Hong Kong/HKPU-5000/2020\|EPI_ISL_610221\|20 | ···················· | ···················· | ···················· | ···················· | NNNNNNNNNNNNNNNNNNNN | NNNNNNNNNNNNNNNNNNNN | 1 |
| N | 4094 hCoV-19/Hong Kong/HKPU-5161/2020\|EPI_ISL_610223\|20 | ···················· | ·A·················· | ···················· | ···················· | NNNNNNNNNNNNNNNNNNNN | NNNNNNNNNNNNNNNNNNNN | 1 |

| | | | | | | | | | |
|---|---|---|---|---|---|---|---|---|---|
| N | 4095 | hCoV-19/Hong Kong/HKPU-5183/2020\|EPI_ISL_610224\|20 | ···················· | ·A·················· | ···················· | ···················· | NNNNNNNNNNNNNNNNNNNN | NNNNNNNNNNNNNNNNNNNN | 1 |
| N | 4519 | hCoV-19/Germany/NW-HHU-192/2020\|EPI_ISL_602466\|202 | ···················· | ···················· | ···················· | ···················· | NNNNNNNNNNNNNNNNNNNN | NNNNNNNNNNNNNNNNNNNN | 1 |
| N | 4542 | hCoV-19/Germany/NW-HHU-168/2020\|EPI_ISL_602490\|202 | ···················· | ···················· | ···················· | ···················· | NNNNNNNNNNNNNNNNNNNN | NNNNNNNNNNNNNNNNNNNN | 1 |
| N | 6148 | hCoV-19/USA/MI-UM-10036307468/2020\|EPI_ISL_596607\| | ···················· | ···················· | ···················· | ···················· | NNNNNNNNNNNNNNNNNNNN | ···················· | 1 |
| N | 6151 | hCoV-19/USA/MI-UM-10036304383/2020\|EPI_ISL_596610\| | ···················· | ···················· | ···················· | ···················· | NNNNNNNNNNNNNNNNNNNN | NNNNNNNNNNNNNNNNNNNN | 1 |
| N | 6152 | hCoV-19/USA/MI-UM-10036369993/2020\|EPI_ISL_596611\| | ···················· | ···················· | ···················· | ···················· | NNNNNNNNNNNNNNNNNNNN | NNNNNNNNNNNNNNNNNNNN | 1 |
| N | 6153 | hCoV-19/USA/MI-UM-10036367756/2020\|EPI_ISL_596612\| | ···················· | ···················· | ···················· | ···················· | NNNNNNNNNNNNNNNNNNNN | NNNNNNNNNNNNNNNNNNNN | 1 |
| N | 6300 | hCoV-19/South Africa/KRISP-K003726/2020\|EPI_ISL_60 | ···················· | ···················· | ···················· | ···················· | ···················· | NNNNNNNNNNNNNNNNNNNN | 1 |
| N | 6301 | hCoV-19/South Africa/KRISP-K003728/2020\|EPI_ISL_60 | ···················· | ···················· | ···················· | ···················· | ·NNNNNNNNNNNNNNNNNN· | ···················· | 1 |
| N | 6311 | hCoV-19/South Africa/KRISP-K003748/2020\|EPI_ISL_60 | ···················· | ···················· | ···················· | ···················· | NNNNNNNNNNNNNNNNNNNN | ···················· | 1 |
| N | 6313 | hCoV-19/South Africa/KRISP-K003750/2020\|EPI_ISL_60 | ···················· | ···················· | ···················· | ···················· | ···················N | NNNNNNNNNNNNNNNNNNNN | 1 |
| N | 6403 | hCoV-19/USA/MI-UM-10036423734/2020\|EPI_ISL_596626\| | ···················· | ···················· | ···················· | ···················· | NNNNNNNNNNNNNNNNNNNN | NNNNNNNNNNNNNNNNNNNN | 1 |
| N | 6632 | hCoV-19/USA/MN-MDH-1821/2020\|EPI_ISL_602956\|2020-0 | ···················· | ·A·················· | ···················· | ···················· | ··NNNNNNNNNN········ | ···················· | 1 |
| N | 6813 | hCoV-19/South Africa/KRISP-K003687/2020\|EPI_ISL_60 | ···················· | ···················· | ···················· | ···················· | ···················· | ·········NNNNNN····· | 1 |
| N | 6820 | hCoV-19/South Africa/KRISP-K003697/2020\|EPI_ISL_60 | ···················· | ···················· | ···················· | ···················· | NNNNNNNNNNNNNNNNNNN· | ···················· | 1 |
| N | 6968 | hCoV-19/USA/MI-UM-10036333501/2020\|EPI_ISL_596584\| | ···················· | ···················· | ···················· | ···················· | NNNNNNNNNNNNNNNNNNNN | ···················· | 1 |
| N | 6970 | hCoV-19/USA/MI-UM-10036312532/2020\|EPI_ISL_596586\| | ···················· | ···················· | ···················· | ···················· | NNNNNNNNNNNNNNNNNNNN | ···················· | 1 |
| N | 7047 | hCoV-19/England/QEUH-9FF35D/2020\|EPI_ISL_600198\|20 | ···················· | NNNNNNNNNNNNNNNNNNNN | ···················· | ···················· | ···················· | ···················· | 1 |
| N | 7127 | hCoV-19/South Africa/KRISP-K003501/2020\|EPI_ISL_60 | ···················· | ···················· | ···················· | ···················· | ···················· | NNNNNNNNNNNNNNNNNNNN | 1 |
| N | 7129 | hCoV-19/South Africa/KRISP-K003504/2020\|EPI_ISL_60 | ···················· | ···················· | ···················· | ···················· | ··NNNNNNNNNNNNNNNNNN | ···················· | 1 |
| N | 7153 | hCoV-19/South Africa/KRISP-K003700/2020\|EPI_ISL_60 | ···················· | ···················· | ···················· | ···················· | NNNNNNNNNNNNNNNNN··· | ···················· | 1 |
| N | 7161 | hCoV-19/South Africa/KRISP-K003715/2020\|EPI_ISL_60 | ···················· | ···················· | ···················· | ···················· | NNNNNNNNNNNNNNNNNNNN | NNNNNNNNNNNNNNNNNNNN | 1 |
| N | 7171 | hCoV-19/USA/WA-S2872/2020\|EPI_ISL_596403\|2020-09-2 | ···················· | ····NNNNNNNNNNNNNNNN | ···················· | ···················· | ···················· | ···················· | 1 |
| N | 7376 | hCoV-19/South Africa/KRISP-K003355/2020\|EPI_ISL_60 | ···················· | ···················· | ···················· | ···················· | ·NNNNN·············· | ···················· | 1 |
| N | 7393 | hCoV-19/South Africa/KRISP-K003376/2020\|EPI_ISL_60 | ···················· | ·A·················· | ···················· | ···················· | NNNNNNNNNNNNNNNNNN·· | ···················· | 1 |
| N | 7398 | hCoV-19/South Africa/KRISP-K003393/2020\|EPI_ISL_60 | ···················· | ···················· | ···················· | ···················· | ·NNNNNNNNNNNNNNNNNNN | NNNNNNNNNNNNNNNNNNNN | 1 |
| N | 7650 | hCoV-19/South Africa/KRISP-K003329/2020\|EPI_ISL_60 | ···················· | ···················· | ···················· | ···················· | NNNNNNNNNNNNN······· | ···················· | 1 |
| N | 7671 | hCoV-19/South Africa/KRISP-K003389/2020\|EPI_ISL_60 | ···················· | ···················· | ···················· | ···················· | ··NNNNNNNNNNNNNNNN·· | ···················· | 1 |
| N | 7858 | hCoV-19/South Africa/KRISP-K003669/2020\|EPI_ISL_60 | ···················· | ···················· | ···················· | ···················· | NNNNN··············· | ···················· | 1 |
| O | 217 | hCoV-19/Wales/PHWC-47F712/2020\|EPI_ISL_612716\|2020 | ···················· | ···················· | ···················· | ···················· | NNNN················ | ···················· | 1 |
| O | 224 | hCoV-19/Wales/PHWC-47F78B/2020\|EPI_ISL_612723\|2020 | ···················· | ···················· | ···················· | ···················· | NNNNNNNNNNNNNNNNNNNN | ···················· | 1 |
| O | 246 | hCoV-19/Wales/PHWC-47FC31/2020\|EPI_ISL_612789\|2020 | ···················· | ···················· | ···················· | ···················· | NNNNNNNNNNNNNNNNNNNN | NNNNNNNNNNNNNNNNNNNN | 1 |
| O | 247 | hCoV-19/Wales/PHWC-47FC40/2020\|EPI_ISL_612790\|2020 | ···················· | ···················· | ···················· | ···················· | NNNNNNNNNNNNNNNNNNNN | NNNNNNNNNNNNNNNNNNNN | 1 |
| O | 362 | hCoV-19/USA/MI-UM-10036399783/2020\|EPI_ISL_610053\| | ···················· | ···················· | ···················· | ···················· | NNNNNNNNNNNNNNNNNNNN | ···················· | 1 |
| O | 364 | hCoV-19/USA/MI-UM-10036329145/2020\|EPI_ISL_610055\| | ···················· | ···················· | ···················· | ···················· | NNNNNNNNNNNNNNNNNNNN | NNNNNNNNNNNNNNNNNNNN | 1 |
| O | 365 | hCoV-19/USA/MI-UM-10036381452/2020\|EPI_ISL_610057\| | ···················· | ···················· | ···················· | ···················· | ······NNNNNNNNNNNNNN | NNNNNNNNNNNNNNNNNNNN | 1 |
| O | 366 | hCoV-19/USA/MI-UM-10036396729/2020\|EPI_ISL_610058\| | ···················· | ···················· | ···················· | ···················· | ······NNNNNNNNNNNNNN | ···················· | 1 |
| O | 375 | hCoV-19/USA/MI-UM-10036422279/2020\|EPI_ISL_610086\| | ···················· | ···················· | ···················· | ···················· | NNNNNNNNNNNNNNNNNNNN | NNNNNNNNNNNNNNNNNNNN | 1 |
| O | 376 | hCoV-19/USA/MI-UM-10036396974/2020\|EPI_ISL_610087\| | ···················· | ···················· | ···················· | ···················· | NNNNNNNNNNNNNNNNNNNN | NNNNNNNNNNNNNNNNNNNN | 1 |
| O | 377 | hCoV-19/USA/MI-UM-10036396723/2020\|EPI_ISL_610088\| | ···················· | ···················· | ···················· | ···················· | NNNNNNNNNNNNNNNNNNNN | ···················· | 1 |
| O | 379 | hCoV-19/USA/MI-UM-10036434968/2020\|EPI_ISL_610090\| | ···················· | ···················· | ···················· | ···················· | NNNNNNNNNNNNNNNNNNNN | ···················· | 1 |
| O | 380 | hCoV-19/USA/MI-UM-10036404933/2020\|EPI_ISL_610092\| | ···················· | ·············A······ | ···················· | ···················· | NNNNNNNNNNNNNNNNNNNN | ···················· | 1 |
| O | 381 | hCoV-19/USA/MI-UM-10036429642/2020\|EPI_ISL_610093\| | ···················· | ···················· | ···················· | ···················· | ·····NNNNNNNNNNNNNNN | ···················· | 1 |
| O | 382 | hCoV-19/USA/MI-UM-10036429659/2020\|EPI_ISL_610094\| | ···················· | ···················· | ···················· | ···················· | NNNNNNNNNNNNNNNNNNNN | NNNNNNNNNNNNNNNNNNNN | 1 |
| O | 384 | hCoV-19/USA/MI-UM-10036421549/2020\|EPI_ISL_610097\| | ···················· | ···················· | ···················· | ···················· | NNNNNNNNNNNNNNNNNNNN | NNNNNNNNNNNNNNNNNNNN | 1 |
| O | 386 | hCoV-19/USA/MI-UM-10036448245/2020\|EPI_ISL_610103\| | ···················· | ···················· | ···················· | ···················· | NNNNNNNNNNNNNNNNNNNN | NNNNNNNNNNNNNNNNNNNN | 1 |
| O | 388 | hCoV-19/USA/MI-UM-10036467524/2020\|EPI_ISL_610105\| | ···················· | ···················· | ···················· | ···················· | NNNNNNNNNNNNNNNNNNNN | NNNNNNNNNNNNNNNNNNNN | 1 |
| O | 389 | hCoV-19/USA/MI-UM-10036444936/2020\|EPI_ISL_610106\| | ···················· | ·············A······ | ···················· | ···················· | NNNNNNNNNNNNNNNNNNNN | NNNNNNNNNNNNNNNNNNNN | 1 |
| O | 390 | hCoV-19/USA/MI-UM-10036449620/2020\|EPI_ISL_610107\| | ···················· | ···················· | ···················· | ···················· | NNNNNNNNNNNNNNNNNNNN | NNNNNNNNNNNNNNNNNNNN | 1 |
| O | 391 | hCoV-19/USA/MI-UM-10036469028/2020\|EPI_ISL_610109\| | ···················· | ···················· | ···················· | ···················· | NNNNNNNNNNNNNNNNNNNN | NNNNNNNNNNNNNNNNNNNN | 1 |
| O | 392 | hCoV-19/USA/MI-UM-10036442334/2020\|EPI_ISL_610110\| | ···················· | ···················· | ···················· | ···················· | NNNNNNNNNNNNNNNNNNNN | ···················· | 1 |
| O | 394 | hCoV-19/USA/MI-UM-10036434093/2020\|EPI_ISL_610112\| | ···················· | ·A·················· | ···················· | ···················· | NNNNNNNNNNNNNNNNNNNN | ···················· | 1 |
| O | 395 | hCoV-19/USA/MI-UM-10036468967/2020\|EPI_ISL_610116\| | ···················· | ···················· | ···················· | ···················· | NNNNNNNNNNNNNNNNNNNN | NNNNNNNNNNNNNNNNNNNN | 1 |
| O | 499 | hCoV-19/Wales/PHWC-47FA82/2020\|EPI_ISL_612765\|2020 | ···················· | ···················· | ···················· | ···················· | NNNNNNNNNNNNNNNNNNNN | NNNNNNNNNNNNNNNNNNNN | 1 |
| O | 509 | hCoV-19/Wales/PHWC-47FB34/2020\|EPI_ISL_612775\|2020 | ···················· | ···················· | ···················· | ···················· | NNNNNNNNNNNNNN······ | ···················· | 1 |
| O | 602 | hCoV-19/USA/MI-UM-10036346836/2020\|EPI_ISL_610061\| | ···················· | ·A·················· | ···················· | ···················· | ······NNNNNNNNNNNNNN | ···················· | 1 |
| O | 603 | hCoV-19/USA/MI-UM-10036386932/2020\|EPI_ISL_610062\| | ···················· | ···················· | ···················· | ···················· | NNNNNNNNNNNNNNNNNNNN | ···················· | 1 |
| O | 605 | hCoV-19/USA/MI-UM-10036381059/2020\|EPI_ISL_610064\| | ···················· | ···················· | ···················· | ···················· | NNNNNNNNNNNNNNNNNNNN | ···················· | 1 |
| O | 607 | hCoV-19/USA/MI-UM-10036347220/2020\|EPI_ISL_610070\| | ···················· | ···················· | ···················· | ···················· | NNNNNNNNNNNNNNNNNNNN | NNNNNNNNNNNNNNNNNNNN | 1 |
| O | 608 | hCoV-19/USA/MI-UM-10036381483/2020\|EPI_ISL_610071\| | ···················· | ···················· | ···················· | ···················· | NNNNNNNNNNNNNNNNNNNN | NNNNNNNNNNNNNNNNNNNN | 1 |
| O | 732 | hCoV-19/USA/FL-BPHL-1824/2020\|EPI_ISL_613911\|2020- | ···················· | ···················· | ···················· | ···················· | NNNNNNNNNNNNNNNNNNNN | ···················· | 1 |
| O | 747 | hCoV-19/USA/FL-BPHL-1846/2020\|EPI_ISL_613926\|2020- | ···················· | ···················· | ···················· | ···················· | NNNNNNNNNNNNNNNNNNNN | ···················· | 1 |
| O | 831 | hCoV-19/England/MILK-A7AB51/2020\|EPI_ISL_606778\|20 | ···················· | ···················· | ···················· | NNNNNNNNNNNNNNNNNNNN | ···················· | ···················· | 1 |
| O | 841 | hCoV-19/England/MILK-A79A73/2020\|EPI_ISL_606789\|20 | ···················· | ···················· | ···················· | NNNNNNNNNNNNNNNNNNNN | ···················· | ···················· | 1 |
| O | 942 | hCoV-19/Northern Ireland/NIRE-108EA3/2020\|EPI_ISL_ | ···················· | ···················· | ···················· | ···················· | NNNNNNNNNNNNNNNNNNNN | NNNNNNNNNNNNNNNNNNNN | 1 |
| O | 951 | hCoV-19/England/NOTT-117779/2020\|EPI_ISL_612586\|20 | ···················· | ···················· | ···················· | ···················· | NNNNNNNNNNNNNNNNNNNN | NNNNNNNNNNNNNNNNNNNN | 1 |
| O | 965 | hCoV-19/Wales/PHWC-47EFDF/2020\|EPI_ISL_612614\|2020 | ···················· | ···················· | ···················· | ···················· | NNNNNNNNNNNNNNNNNNNN | ···················· | 1 |
| O | 973 | hCoV-19/Wales/PHWC-47F0CC/2020\|EPI_ISL_612626\|2020 | ···················· | ···················· | ···················· | ···················· | ···················· | NNNNNNNNNNNNNNN····· | 1 |
| O | 1007 | hCoV-19/USA/FL-BPHL-1763/2020\|EPI_ISL_613870\|2020- | ···················· | ···················· | ···················· | ···················· | NNNNNNNNNNNNNNNNNNNN | ···················· | 1 |
| O | 1009 | hCoV-19/USA/FL-BPHL-1766/2020\|EPI_ISL_613872\|2020- | ···················· | ···················· | ···················· | ···················· | NNNNNNNNNNNNNNNNNNNN | ···················· | 1 |
| O | 1010 | hCoV-19/USA/FL-BPHL-1768/2020\|EPI_ISL_613873\|2020- | ···················· | ···················· | ···················· | ···················· | NNNNNNNNNNNNNNNNNNNN | ···················· | 1 |
| O | 1171 | hCoV-19/England/MILK-A193A7/2020\|EPI_ISL_612509\|20 | ···················· | ···················· | ···················· | ···················· | NNNNNNNNNNNNNNNNNNNN | NNNNN··············· | 1 |
| O | 1178 | hCoV-19/England/MILK-A19547/2020\|EPI_ISL_612516\|20 | ···················· | ···················· | ···················· | ···················· | NNNNNNNNNNNNNNNNNNNN | NNNNNNNNNNNNNNNNNNNN | 1 |
| O | 1195 | hCoV-19/England/MILK-A229A9/2020\|EPI_ISL_612533\|20 | ···················· | ···················· | ···················· | ···················· | NNNNNNNNNNNNNNNNNNNN | NNNNNNNNNNNNNNNNNNNN | 1 |
| O | 1233 | hCoV-19/USA/FL-BPHL-1905/2020\|EPI_ISL_613832\|2020- | ···················· | ···················· | ···················· | ···················· | NNNNNNNNNNNNNNNNNNNN | NNNNNNNNNNNNNNNNNNNN | 1 |
| O | 1234 | hCoV-19/USA/FL-BPHL-1906/2020\|EPI_ISL_613833\|2020- | ···················· | ···················· | ···················· | ···················· | NNNNNNNNNNNNNNNNNNNN | NNNNNNNNNNNNNNNNNNNN | 1 |
| O | 1238 | hCoV-19/USA/FL-BPHL-1710/2020\|EPI_ISL_613844\|2020- | ···················· | ·············A······ | ···················· | ···················· | NNNNNNNNNNNNNNNNNNNN | ···················· | 1 |
| O | 1239 | hCoV-19/USA/FL-BPHL-1712/2020\|EPI_ISL_613845\|2020- | ···················· | ·············A······ | ···················· | ···················· | NNNN················ | ···················· | 1 |
| O | 1242 | hCoV-19/USA/FL-BPHL-1717/2020\|EPI_ISL_613848\|2020- | ···················· | ···················· | ···················· | ···················· | NNNN················ | ···················· | 1 |
| O | 1254 | hCoV-19/USA/FL-BPHL-1742/2020\|EPI_ISL_613860\|2020- | ···················· | ·············A······ | ···················· | ···················· | NNNNNNNNNNNNNNNNNNNN | ···················· | 1 |

| | | | | | | | | | |
|---|---|---|---|---|---|---|---|---|---|
| O | 1273 | hCoV-19/USA/FL-BPHL-1790/2020\|EPI_ISL_613892\|2020- | ···················· | ···················· | ···················· | ···················· | NNNNNNNNNNNNNNNNNNNN | ···················· | 1 |
| O | 1280 | hCoV-19/USA/FL-BPHL-1800/2020\|EPI_ISL_613899\|2020- | ···················· | ···················· | ···················· | ···················· | NNNNNNNNNNNNNNNNNNNN | NNNNNNNNNNNNNNNNNNNN | 1 |
| O | 1511 | hCoV-19/England/MILK-A190A0/2020\|EPI_ISL_612480\|20 | ···················· | ···················· | ···················· | ···················· | NNNNNNNNNNNNNNNNNNNN | NNNNNNNNNNNNNNNNNNNN | 1 |
| O | 1516 | hCoV-19/Scotland/MILK-A19134/2020\|EPI_ISL_612485\|2 | ···················· | ···················· | ···················· | ···················· | NNNN················ | ···················· | 1 |
| O | 1529 | hCoV-19/England/MILK-A192C8/2020\|EPI_ISL_612498\|20 | ···················· | ·A·················· | ···················· | ···················· | NNNNNNNNNNNNNNNNNNNN | ···A················ | 1 |
| O | 1534 | hCoV-19/USA/FL-BPHL-1895/2020\|EPI_ISL_613769\|2020- | ···················· | ···················· | ···················· | ···················· | ·······NNNNNNNNNNNNN | ···················· | 1 |
| O | 1691 | hCoV-19/England/LIVE-1D2A80/2020\|EPI_ISL_612404\|20 | ···················· | ···················· | ···················· | ···················· | NNNNNNNNNNNNNNNNNNNN | ···················· | 1 |
| O | 1693 | hCoV-19/England/LIVE-1D2ACC/2020\|EPI_ISL_612406\|20 | ···················· | ···················· | ···················· | ···················· | NNNN···NNNNNNNNNNNNN | ···················· | 1 |
| O | 1701 | hCoV-19/England/LIVE-1D2BC9/2020\|EPI_ISL_612414\|20 | ···················· | ···················· | ···················· | ···················· | NNNNNNNNNNNNNNNNNNNN | NNNNNNNNNNNNNNNNNNNN | 1 |
| O | 1778 | hCoV-19/USA/MI-MDHHS-SC22115/2020\|EPI_ISL_614166\|2 | ···················· | ···················· | ···················· | ···················· | NNNNNNNNNNNNNNNNNNNN | ···················· | 1 |
| O | 1780 | hCoV-19/USA/MI-MDHHS-SC22118/2020\|EPI_ISL_614169\|2 | ···················· | ·A·················· | ···················· | ···················· | NNNNNNNNNNNNNNNNNNNN | ···················· | 1 |
| O | 1781 | hCoV-19/USA/MI-MDHHS-SC22119/2020\|EPI_ISL_614170\|2 | ···················· | ·A·················· | ···················· | ···················· | NNNNNNNNNNNNNNNNNNNN | ···················· | 1 |
| O | 1782 | hCoV-19/USA/MI-MDHHS-SC22120/2020\|EPI_ISL_614171\|2 | ···················· | ·A·················· | ···················· | ···················· | NNNNNNNNNNNNNNNNNNNN | ···················· | 1 |
| O | 1834 | hCoV-19/England/ALDP-A86E1D/2020\|EPI_ISL_605983\|20 | ···················· | NNNNNNNNNNNNNNNNNNNN | ···················· | ···················· | ···················· | ···················· | 1 |
| O | 2492 | hCoV-19/Wales/PHWC-48063F/2020\|EPI_ISL_612913\|2020 | ···················· | ···················· | ···················· | ···················· | NNNNNNNNNNNNN······· | ···················· | 1 |
| O | 2512 | hCoV-19/Wales/PHWC-4807E1/2020\|EPI_ISL_612933\|2020 | ···················· | ···················· | ···················· | ···················· | NNNNNNNNNNNNNNNNNNNN | ·················NNN | 1 |
| O | 2515 | hCoV-19/Wales/PHWC-48081B/2020\|EPI_ISL_612936\|2020 | ···················· | ···················· | ···················· | ···················· | ···················· | ·····NNNNNNNNNN····· | 1 |
| O | 2519 | hCoV-19/Wales/PHWC-480857/2020\|EPI_ISL_612940\|2020 | ···················· | ···················· | ···················· | ···················· | NNNNNNNNNNNNNNNNNNNN | ················NNNN | 1 |
| O | 2536 | hCoV-19/Wales/PHWC-4809AF/2020\|EPI_ISL_612957\|2020 | ···················· | ···················· | ···················· | ···················· | ···NNNNNNNNNNNNNNNNN | NNNNNNNNNNNNNNN····· | 1 |
| O | 2548 | hCoV-19/Wales/PHWC-480AD9/2020\|EPI_ISL_612969\|2020 | ···················· | ···················· | ···················· | ···················· | NNNNNNNNNNNNNNNNNNNN | ···················· | 1 |
| O | 2554 | hCoV-19/Wales/PHWC-480B4F/2020\|EPI_ISL_612975\|2020 | ···················· | ···················· | ···················· | ···················· | ·············NNNNNNN | NNNNNNNNNNNNNNNNNNNN | 1 |
| O | 2555 | hCoV-19/Wales/PHWC-480B5E/2020\|EPI_ISL_612976\|2020 | ···················· | ···················· | ···················· | ···················· | ···················· | NNNNNNNN···NNNN····· | 1 |
| O | 2768 | hCoV-19/Wales/PHWC-47FEC2/2020\|EPI_ISL_612825\|2020 | ···················· | ···················· | ···················· | ···················· | NNNNNNNNNNNNNNNNNNNN | NNNNNNNNNNNNNNNNNNNN | 1 |
| O | 2788 | hCoV-19/Wales/PHWC-48006E/2020\|EPI_ISL_612845\|2020 | ···················· | ···················· | ···················· | ···················· | NNNNNNNNNNNNNNNNNNNN | ···················· | 1 |
| O | 2791 | hCoV-19/Wales/PHWC-4800AA/2020\|EPI_ISL_612848\|2020 | ···················· | ···················· | ···················· | ···················· | NNNNNNNNNNNNNNNNNNNN | NNNNNNNNNNNNNNNNNNNN | 1 |
| O | 2792 | hCoV-19/Wales/PHWC-4800B9/2020\|EPI_ISL_612849\|2020 | ···················· | ···················· | ···················· | ···················· | ···················· | NNNN················ | 1 |
| O | 2822 | hCoV-19/England/MILK-A7BA9F/2020\|EPI_ISL_607405\|20 | ···················· | NNNNNNNNNNNNNNNNNNNN | ···················· | ···················· | ···················· | ···················· | 1 |
| O | 2952 | hCoV-19/Wales/PHWC-480268/2020\|EPI_ISL_612867\|2020 | ···················· | ···················· | ···················· | ···················· | NNNNNNNNNNNNNNNNNNNN | NNNNNNNNNNNNNNNNNNN· | 1 |
| O | 2953 | hCoV-19/Wales/PHWC-480277/2020\|EPI_ISL_612868\|2020 | ···················· | ···················· | ···················· | ···················· | NNNNNNNNNNNNNNNNNNNN | ···NNNNNNNNNNN······ | 1 |
| O | 2985 | hCoV-19/Wales/PHWC-48212D/2020\|EPI_ISL_613200\|2020 | ···················· | ···················· | ···················· | ···················· | ·······NNNNNNNNNNNNN | NNNNNNNNN········NNN | 1 |
| O | 3018 | hCoV-19/Wales/PHWC-48261F/2020\|EPI_ISL_613233\|2020 | ···················· | ···················· | ···················· | ···················· | NNNNNNNNNNNNNNNNNNNN | ···················· | 1 |
| O | 3020 | hCoV-19/Wales/PHWC-48265B/2020\|EPI_ISL_613235\|2020 | ···················· | ···················· | ···················· | ···················· | NNNNNNNNNNNNNNNNNNNN | ···················· | 1 |
| O | 3027 | hCoV-19/Wales/PHWC-4827A3/2020\|EPI_ISL_613242\|2020 | ···················· | ···················· | ···················· | ···················· | ·NNNNNNNNNNNNNNNNNNN | ···················· | 1 |
| O | 3035 | hCoV-19/Wales/PHWC-482882/2020\|EPI_ISL_613250\|2020 | ···················· | ···················· | ···················· | ···················· | ···················· | NNNNNNNNNNNNNNN····· | 1 |
| O | 3036 | hCoV-19/Wales/PHWC-4828DD/2020\|EPI_ISL_613251\|2020 | ···················· | ···················· | ···················· | ···················· | NNNNNNNNNNNNNNNNNNNN | NNNNNNNNNNNNNNNNNNNN | 1 |
| O | 3039 | hCoV-19/Wales/PHWC-482952/2020\|EPI_ISL_613254\|2020 | ···················· | ···················· | ···················· | ···················· | NNNNNNNNNNNNNNNNNNNN | ···················· | 1 |
| O | 3041 | hCoV-19/Wales/PHWC-4829CB/2020\|EPI_ISL_613256\|2020 | ···················· | ···················· | ···················· | ···················· | ···················· | NNNNNNNN············ | 1 |
| O | 3165 | hCoV-19/England/MILK-A7CA07/2020\|EPI_ISL_607368\|20 | ···················· | ···················· | ···················· | NNNNNNNNNNNNNNNNNNNN | ···················· | ···················· | 1 |
| O | 3276 | hCoV-19/Italy/LAZ-INMI-88/2020\|EPI_ISL_609996\|2020 | ···················· | ···················· | ···················· | ···················· | NNNNNNNNNNNNNNN····· | ···················· | 1 |
| O | 3277 | hCoV-19/Italy/LAZ-INMI-89/2020\|EPI_ISL_609997\|2020 | ···················· | ···················· | ···················· | ···················· | NNNNNNNNNNNNNNN····· | ···················· | 1 |
| O | 3485 | hCoV-19/Wales/PHWC-4816C5/2020\|EPI_ISL_613100\|2020 | ···················· | ···················· | ···················· | ···················· | NNNNNNNNNNNNNNNNNNNN | NNNNNNNNNNNNNNNNNNNN | 1 |
| O | 3488 | hCoV-19/Wales/PHWC-48174A/2020\|EPI_ISL_613103\|2020 | ···················· | ···················· | ···················· | ···················· | NNNNNNNNNNNNNNNNNNNN | NNNNNNNNNNNNNNNNNNNN | 1 |
| O | 3489 | hCoV-19/Wales/PHWC-481759/2020\|EPI_ISL_613104\|2020 | ···················· | ···················· | ···················· | ···················· | NNNNNNNNNNNNNNNNNNNN | NNNNNNNNNNNNNNNNNNNN | 1 |
| O | 3492 | hCoV-19/Wales/PHWC-481795/2020\|EPI_ISL_613107\|2020 | ···················· | ···················· | ···················· | ···················· | NNNNNNNNNNNNNNNNNNNN | ···················· | 1 |
| O | 3501 | hCoV-19/Wales/PHWC-481838/2020\|EPI_ISL_613116\|2020 | ···················· | ···················· | ···················· | ···················· | ················NNNN | ···················· | 1 |
| O | 3502 | hCoV-19/Wales/PHWC-481847/2020\|EPI_ISL_613117\|2020 | ···················· | ···················· | ···················· | ···················· | NNNNNNNNNNNNNNNNNNNN | NNNNNNNNNNNNNNNNNNNN | 1 |
| O | 3514 | hCoV-19/Wales/PHWC-481980/2020\|EPI_ISL_613129\|2020 | ···················· | ···················· | ···················· | ···················· | NNNNNNNNNNNNNNNNNNNN | ···················· | 1 |
| O | 3524 | hCoV-19/Wales/PHWC-481A8D/2020\|EPI_ISL_613139\|2020 | ···················· | ···················· | ···················· | ···················· | ···················· | NNNNN··············· | 1 |
| O | 3526 | hCoV-19/Wales/PHWC-481ABA/2020\|EPI_ISL_613141\|2020 | ···················· | ···················· | ···················· | ···················· | NNNNNNNNNNNNNNNNNNNN | NNNNNNNNNNNNNNNNNNNN | 1 |
| O | 3529 | hCoV-19/Wales/PHWC-481AE7/2020\|EPI_ISL_613144\|2020 | ···················· | ···················· | ···················· | ···················· | ···················· | NNNNNNNN············ | 1 |
| O | 3531 | hCoV-19/Wales/PHWC-481B11/2020\|EPI_ISL_613146\|2020 | ···················· | ···················· | ···················· | ···················· | ··NNNNNNNNNNNNNNNNNN | ···················· | 1 |
| O | 3532 | hCoV-19/Wales/PHWC-481B4E/2020\|EPI_ISL_613147\|2020 | ···················· | ···················· | ···················· | ···················· | ···················· | N········NNNNNN··NNN | 1 |
| O | 3535 | hCoV-19/Wales/PHWC-481B8A/2020\|EPI_ISL_613150\|2020 | ···················· | ···················· | ···················· | ···················· | NNNNNNNNNNNNNNNNNNNN | NNNNNNNNNNNNNNNNNNNN | 1 |
| O | 3536 | hCoV-19/Wales/PHWC-481B99/2020\|EPI_ISL_613151\|2020 | ···················· | ···················· | ···················· | ···················· | NNNNNNNNNNNNNNNNNNNN | NNNNNNNNNNNNNNNNNNNN | 1 |
| O | 3544 | hCoV-19/Wales/PHWC-481C4B/2020\|EPI_ISL_613159\|2020 | ···················· | ···················· | ···················· | ···················· | ···················· | NNNNNNNNNNNNNNN····· | 1 |
| O | 3548 | hCoV-19/Wales/PHWC-481CD2/2020\|EPI_ISL_613163\|2020 | ···················· | ···················· | ···················· | ···················· | NNNNNNNNNNNNNNNNNNNN | NNNNNNNNNNNNNNNNNNNN | 1 |
| O | 3566 | hCoV-19/Wales/PHWC-481E72/2020\|EPI_ISL_613181\|2020 | ···················· | ···················· | ···················· | ···················· | NNNNNNNNNNNNNNNNNNNN | ···················· | 1 |
| O | 3577 | hCoV-19/Wales/PHWC-481FE8/2020\|EPI_ISL_613192\|2020 | ···················· | ···················· | ···················· | ···················· | ···················· | NNNNNNNNN··········· | 1 |
| O | 4007 | hCoV-19/Wales/PHWC-480E73/2020\|EPI_ISL_613010\|2020 | ···················· | ···················· | ···················· | ···················· | NNNNNNNNNNNNNNNNNNNN | NNNNNNNNNNNNNNNNNNNN | 1 |
| O | 4010 | hCoV-19/Wales/PHWC-480EFB/2020\|EPI_ISL_613013\|2020 | ···················· | ···················· | ···················· | ···················· | NNNNNNNNNNNNNNNNNNNN | NNNNNNNNNNNNNNNNNNNN | 1 |
| O | 4011 | hCoV-19/Wales/PHWC-480F07/2020\|EPI_ISL_613014\|2020 | ···················· | ···················· | ···················· | ···················· | NNNNNNNNNNNNNNNNNNNN | ···················· | 1 |
| O | 4012 | hCoV-19/Wales/PHWC-480F16/2020\|EPI_ISL_613015\|2020 | ···················· | ···················· | ···················· | ···················· | NNNNNNNNNNNN········ | ···················· | 1 |
| O | 4019 | hCoV-19/Wales/PHWC-480FAD/2020\|EPI_ISL_613022\|2020 | ···················· | ···················· | ···················· | ···················· | ·NNNNNNNNNNN········ | ···················· | 1 |
| O | 4020 | hCoV-19/Wales/PHWC-480FCB/2020\|EPI_ISL_613023\|2020 | ···················· | ···················· | ···················· | ···················· | NNNNNNNNNNNNNNNNNNNN | NNNNNNNNNNNNNNNNNNNN | 1 |
| O | 4022 | hCoV-19/Wales/PHWC-480FE9/2020\|EPI_ISL_613025\|2020 | ···················· | ···················· | ···················· | ···················· | NNNNNNNNNNNNNNNNNNNN | ···················· | 1 |
| O | 4026 | hCoV-19/Wales/PHWC-48105E/2020\|EPI_ISL_613029\|2020 | ···················· | ···················· | ···················· | ···················· | NNNNNNNNNNNNNNNNNNNN | NNNNNNNNNNNNNNNNNNNN | 1 |
| O | 4027 | hCoV-19/Wales/PHWC-48106D/2020\|EPI_ISL_613030\|2020 | ···················· | ···················· | ···················· | ···················· | NNNNNNNNNNNNNNNNNNNN | NNNNNNNNNNNNNNN····· | 1 |
| O | 4028 | hCoV-19/Wales/PHWC-48107C/2020\|EPI_ISL_613031\|2020 | ···················· | ···················· | ···················· | ···················· | NNNNNNNNNNNNNNNNNNNN | ···················· | 1 |
| O | 4032 | hCoV-19/Wales/PHWC-48111F/2020\|EPI_ISL_613035\|2020 | ···················· | ···················· | ···················· | ···················· | NNNNNNNNNNNNNNNNNNNN | NNNNNNNNNNNNNNNNNNNN | 1 |
| O | 4041 | hCoV-19/Wales/PHWC-4811E2/2020\|EPI_ISL_613044\|2020 | ···················· | ···················· | ···················· | ···················· | NNNNNNNNNNNNNNNNNNNN | NNNNNNNNNNNNNNNNNNNN | 1 |
| O | 4047 | hCoV-19/Wales/PHWC-481285/2020\|EPI_ISL_613050\|2020 | ···················· | ···················· | ···················· | ···················· | ···················· | NNNNNNNNNNN········· | 1 |
| O | 4050 | hCoV-19/Wales/PHWC-4812EF/2020\|EPI_ISL_613053\|2020 | ···················· | ···················· | ···················· | ···················· | NNNNNNNNNNNNNNNNNNNN | NNNNNNNNNNNNNNN····· | 1 |
| O | 4051 | hCoV-19/Wales/PHWC-48130A/2020\|EPI_ISL_613054\|2020 | ···················· | ···················· | ···················· | ···················· | NNNNNNNNNNNNNNNNNNNN | NNNNNNNNNNNNNNNNNNNN | 1 |
| O | 4053 | hCoV-19/Wales/PHWC-481337/2020\|EPI_ISL_613056\|2020 | ···················· | ···················· | ···················· | ···················· | NNNNNNNNNNNNNNNNNNNN | NNNNNNNNNNNNNNNNNNNN | 1 |
| O | 4055 | hCoV-19/Wales/PHWC-481355/2020\|EPI_ISL_613058\|2020 | ···················· | ···················· | ···················· | ···················· | NNNNNNNNNNNNNNNNNNNN | NNNNNNNNNNNNNNNNNNNN | 1 |
| O | 4059 | hCoV-19/Wales/PHWC-4813A0/2020\|EPI_ISL_613062\|2020 | ···················· | ···················· | ···················· | ···················· | NNNNNNNNNNNNNNNNNNNN | NNNNNNNNNNNNNNNNNNNN | 1 |
| O | 4063 | hCoV-19/Wales/PHWC-4813FB/2020\|EPI_ISL_613066\|2020 | ···················· | ···················· | ···················· | ···················· | NNNNNNNNNNNNNNNNNNNN | NNNNNNNNNNNNNNNNNNNN | 1 |
| O | 4067 | hCoV-19/Wales/PHWC-481443/2020\|EPI_ISL_613070\|2020 | ···················· | ···················· | ···················· | ···················· | NNNNNNNNNNNNNNNNNNNN | NNNNNNNNNNNNNNNNNNNN | 1 |
| O | 4068 | hCoV-19/Wales/PHWC-481461/2020\|EPI_ISL_613071\|2020 | ···················· | ···················· | ···················· | ···················· | NNNNNNNNNNNNNNNNNNNN | NNNNNNNNNNNNNNNNNNNN | 1 |
| O | 4069 | hCoV-19/Wales/PHWC-481470/2020\|EPI_ISL_613072\|2020 | ···················· | ···················· | ···················· | ···················· | NNNNNNNNNNNNNNNNNNNN | NNNNNNNNNNNNNNNNNNNN | 1 |

| | | | | | | | | |
|---|---|---|---|---|---|---|---|---|
| O | 4071 hCoV-19/Wales/PHWC-48149E/2020\|EPI_ISL_613074\|2020 | ···················· | ···················· | ···················· | ···················· | ···················· | NNNNNNNNNNNNNNN····· | 1 |
| O | 4075 hCoV-19/Wales/PHWC-4814F8/2020\|EPI_ISL_613078\|2020 | ···················· | ···················· | ···················· | ···················· | NNNNNNNNNNNNNNNNNNNN | NNNNNNNNNNNNNNNNNNNN | 1 |
| O | 4082 hCoV-19/Wales/PHWC-48156E/2020\|EPI_ISL_613085\|2020 | ···················· | ···················· | ···················· | ···················· | NNNNNNNNNNNNNNNNNNNN | ···················· | 1 |
| O | 4091 hCoV-19/Wales/PHWC-48162F/2020\|EPI_ISL_613094\|2020 | ···················· | ···················· | ···················· | ···················· | NNNNNNNNNNNNNNNNNNNN | NNNNNNNNNNNNNNNNNNNN | 1 |
| O | 4093 hCoV-19/Wales/PHWC-48164D/2020\|EPI_ISL_613096\|2020 | ···················· | ···················· | ···················· | ···················· | NNNNNNNNNNNNNNNNNNNN | ···················· | 1 |
| O | 4095 hCoV-19/Wales/PHWC-481689/2020\|EPI_ISL_613098\|2020 | ···················· | ···················· | ···················· | ···················· | NNNNNNNNNNNNNNNNNNNN | NNNNNNNNNNNNNNNNNNNN | 1 |
| O | 4855 hCoV-19/England/EXET-1381FC/2020\|EPI_ISL_612390\|20 | ···················· | ···················· | ···················· | ···················· | NNNNNNNNNNNNNNNNNNNN | NNNNNNNNNNNNNNNNNNNN | 1 |
| O | 4862 hCoV-19/England/EXET-13839C/2020\|EPI_ISL_612397\|20 | ···················· | ···················· | ···················· | ···················· | NNNNNNNNNNNNNNNNNNNN | NNNNNNNNNNNNNNNNNNNN | 1 |
| O | 4864 hCoV-19/England/EXET-13846C/2020\|EPI_ISL_612399\|20 | ···················· | NNNNNNNNNNNNNNNNNNNN | ···················· | ···················· | ···················· | ···················· | 1 |
| O | 5063 hCoV-19/Scotland/EDB8403/2020\|EPI_ISL_612352\|2020- | ···················· | ···················· | ···················· | ···················· | NNNNNNNNNNNNNNNNNNNN | NNNNNNNNNNNNNNNNNNNN | 1 |
| O | 5066 hCoV-19/Scotland/EDB8410/2020\|EPI_ISL_612355\|2020- | ···················· | ···················· | ···················· | ···················· | NNNNNNNNNNNNNNNNNNNN | NNNNNNNNNNNNNNNNNNNN | 1 |
| O | 5079 hCoV-19/Scotland/EDB8473/2020\|EPI_ISL_612368\|2020- | ···················· | ···················· | ···················· | ···················· | NNNNNNNNNNNNNNNNNNNN | NNNNNNNNNNNNNNNNNNNN | 1 |
| O | 5200 hCoV-19/England/BIRM-65D20/2020\|EPI_ISL_612221\|202 | ···················· | ·A·················· | ···················· | ···················· | NNNNNNNNNNNNNNNNNNNN | NNNNNNNNNNNNNNNNNNNN | 1 |
| O | 5207 hCoV-19/England/BIRM-65DB7/2020\|EPI_ISL_612228\|202 | ···················· | ···················· | ···················· | ···················· | NNNNNNNNNNNNNNNNNNNN | NNNNNNNNNNNNNNNNNNNN | 1 |
| O | 5214 hCoV-19/England/CAMB-1B6A55/2020\|EPI_ISL_612266\|20 | ···················· | ···················· | ···················· | ···················· | NNNNNNNNNNNNNNNNNNNN | NNNNNNNNNNNNNNNNNNNN | 1 |
| O | 5252 hCoV-19/Scotland/CVR4668/2020\|EPI_ISL_612304\|2020- | ···················· | ···················· | ···················· | ···················· | NNNNNNNNNNNNNNNNNNNN | NNNNNNNNNNNNNNNNNNNN | 1 |
| O | 5261 hCoV-19/Scotland/CVR4692/2020\|EPI_ISL_612313\|2020- | ···················· | ···················· | ···················· | ···················· | ···················· | NN·N···NN··········· | 1 |
| O | 5576 hCoV-19/England/BIRM-65CBA/2020\|EPI_ISL_612218\|202 | ···················· | ···················· | ···················· | ···················· | NNNNNNNNNNNNNNNNNNNN | NNNNNNNNNNNNNNNNNNNN | 1 |
| O | 5577 hCoV-19/England/BIRM-65CC9/2020\|EPI_ISL_612219\|202 | ···················· | ·A·················· | ···················· | ···················· | NNNNNNNNNNNNNNNNNNNN | NNNNNNNNNNNNNNNNNNNN | 1 |
| O | 5584 hCoV-19/England/BIRM-65FOC/2020\|EPI_ISL_612235\|202 | ···················· | ···················· | ···················· | ···················· | NNNNNNNNNNNNNNNNNNNN | NNNNNNNNNNNNNNNNNNNN | 1 |
| O | 5608 hCoV-19/England/BIRM-66387/2020\|EPI_ISL_612259\|202 | ···················· | ···················· | ···················· | ···················· | NNNNNNNNNNNNNNNNNNNN | NNNNNNNNNNNNNNNNNNNN | 1 |
| O | 5756 hCoV-19/Wales/PHWC-4826F1/2020\|EPI_ISL_612157\|2020 | ···················· | ···················· | ···················· | ···················· | NNNNNNNNNNNNNNNNNNNN | ···················· | 1 |
| O | 5758 hCoV-19/Wales/PHWC-48059C/2020\|EPI_ISL_612203\|2020 | ···················· | ···················· | ···················· | ···················· | N·······NNNNNNNNNNNN | NNNNNNNNNNNNNNNNNNNN | 1 |
| O | 5763 hCoV-19/England/BIRM-65B53/2020\|EPI_ISL_612208\|202 | ···················· | ···················· | ···················· | ···················· | NNNNNNNNNNNNNNNNNNNN | NNNNNNNNNNNNNNNNNNNN | 1 |
| O | 5765 hCoV-19/England/BIRM-65BAE/2020\|EPI_ISL_612210\|202 | ···················· | ···················· | ···················· | ···················· | NNNNNNNNNNNNNNNNNNNN | NNNNNNNNNNNNNNNNNNNN | 1 |
| O | 5766 hCoV-19/England/BIRM-65BBD/2020\|EPI_ISL_612211\|202 | ···················· | ···················· | ···················· | ···················· | NNNNNNNNNNNNNNNNNNNN | NNNNNNNNNNNNNNNNNNNN | 1 |
| O | 5767 hCoV-19/England/BIRM-65BDB/2020\|EPI_ISL_612212\|202 | ···················· | ·A·················· | ···················· | ···················· | NNNNNNNNNNNNNNNNNNNN | NNNNNNNNNNNNNNNNNNNN | 1 |
| O | 5886 hCoV-19/France/BRE-BR9056/2020\|EPI_ISL_613546\|2020 | ···················· | ·A·················· | ···················· | ···················· | NNNNNNNNNNNNNNNNNNNN | ···················· | 1 |
| O | 6109 hCoV-19/Scotland/EDB8429/2020\|EPI_ISL_612188\|2020- | ···················· | ···················· | ···················· | ···················· | NNNNNNNNNNNNNNNNNNNN | NNNNNNNNNNNNNNNNNNNN | 1 |
| O | 6115 hCoV-19/Wales/PHWC-48165C/2020\|EPI_ISL_612194\|2020 | ···················· | ···················· | ···················· | ···················· | ···················· | NNNNNNNNNNNNNNN····· | 1 |
| O | 6172 hCoV-19/England/ALDP-A878A5/2020\|EPI_ISL_606208\|20 | ···················· | ···················· | ···················· | NNNNNNNNNNNNNNNNNNNN | ···················· | ···················· | 1 |
| O | 6188 hCoV-19/England/MILK-A7B006/2020\|EPI_ISL_607475\|20 | ···················· | NNNNNNNNNNNNNNNNNNNN | ···················· | ···················· | ···················· | ···················· | 1 |
| O | 6195 hCoV-19/England/MILK-A7BE39/2020\|EPI_ISL_607483\|20 | ···················· | ···················· | ···················· | NNNNNNNNNNNNNNNNNNNN | ···················· | ···················· | 1 |
| O | 6487 hCoV-19/England/MILK-A7B22E/2020\|EPI_ISL_607457\|20 | ···················· | ···················· | ···················· | NNNNNNNNNNNNNNNNNNN· | ···················· | ···················· | 1 |
| P | 1062 hCoV-19/England/MILK-AB73BA/2020\|EPI_ISL_623239\|20 | ···················· | ···················· | ···················· | ···················· | NNNNNNNNNNNNNNNNNNNN | NNNNNNNNNNNNNNNNNNNN | 1 |
| P | 1959 hCoV-19/Brazil/RJ-00326/2020\|EPI_ISL_623129\|2020-0 | ···················· | ···················· | ···················· | ···················· | ······NNNNNNNNNNNNNN | NNNNNNNNNNNNNNNNNNNN | 1 |
| P | 2443 hCoV-19/Denmark/DCGC-4621/2020\|EPI_ISL_621290\|2020 | ···················· | ···················· | ···················· | ···················· | NNNNNNNNNNNNNNNNNNNN | NNNNNNNNNNNNNNNNNNNN | 1 |
| P | 3160 hCoV-19/Denmark/DCGC-2838/2020\|EPI_ISL_616665\|2020 | ···················· | ···················· | ···················· | ···················· | NNNNNNNNNNNNNNNNNNNN | ···················· | 1 |
| P | 5202 hCoV-19/Denmark/DCGC-8483/2020\|EPI_ISL_618046\|2020 | ···················· | ···················· | ···················· | ···················· | NNNNNNNNNNNNNNNNNNNN | NNNNNNNNNNNNNNNNNNNN | 1 |
| P | 5215 hCoV-19/Denmark/DCGC-8484/2020\|EPI_ISL_618059\|2020 | ···················· | ···················· | ···················· | ···················· | NNNNNNNNNNNNNNNNNNNN | NNNNNNNNNNNNNNNNNNNN | 1 |
| P | 5217 hCoV-19/Denmark/DCGC-8487/2020\|EPI_ISL_618063\|2020 | ···················· | ···················· | ···················· | ···················· | NNNNNNNNNNNNNNNNNNNN | NNNNNNNNNNNNNNNNNNNN | 1 |
| P | 6115 hCoV-19/Belgium/ULG-10547/2020\|EPI_ISL_626305\|2020 | ···················· | ···················· | ···················· | ···················· | NNNNNNNNNNNNNNNNNNNN | NNNNNNNNNNNNNNNNNNNN | 1 |
| P | 6118 hCoV-19/Belgium/ULG-10534/2020\|EPI_ISL_626310\|2020 | ···················· | ···················· | ···················· | ···················· | NNNNNNNNNNNNNNNNNNNN | NNNNNNNNNNNNNNNNNNNN | 1 |
| P | 6125 hCoV-19/Belgium/ULG-10575/2020\|EPI_ISL_626319\|2020 | ···················· | ···················· | ···················· | ···················· | NNNNNNNNNNNNNNNNNNNN | NNNNNNNNNNNNNNNNNNNN | 1 |
| P | 6129 hCoV-19/Belgium/ULG-10562/2020\|EPI_ISL_626323\|2020 | ···················· | ···················· | ···················· | ···················· | NNNNNNNNNNNNNNNNNNNN | NNNNNNNNNNNNNNNNNNNN | 1 |
| P | 6130 hCoV-19/Belgium/ULG-10579/2020\|EPI_ISL_626324\|2020 | ···················· | ···················· | ···················· | ···················· | NNNNNNNNNNNNNNNNNNNN | NNNNNNNNNNNNNNNNNNNN | 1 |
| P | 6135 hCoV-19/Belgium/ULG-10561/2020\|EPI_ISL_626330\|2020 | ···················· | ···················· | ···················· | ···················· | NNNNNNNNNNNNNNNNNNNN | NNNNNNNNNNNNNNNNNNNN | 1 |
| P | 6137 hCoV-19/Belgium/ULG-10563/2020\|EPI_ISL_626332\|2020 | ···················· | ·A·················· | ···················· | ···················· | NNNNNNNNNNNNNNNNNNNN | NNNNNNNNNNNNNNNNNNNN | 1 |
| P | 6138 hCoV-19/Belgium/ULG-10577/2020\|EPI_ISL_626333\|2020 | ···················· | ···················· | ···················· | ···················· | NNNNNNNNNNNNNNNNNNNN | NNNNNNNNNNNNNNNNNNNN | 1 |
| P | 6140 hCoV-19/Belgium/ULG-10595/2020\|EPI_ISL_626335\|2020 | ···················· | ···················· | ···················· | ···················· | NNNNNNNNNNNNNNNNNNNN | NNNNNNNNNNNNNNNNNNNN | 1 |
| P | 6142 hCoV-19/Belgium/ULG-10591/2020\|EPI_ISL_626337\|2020 | ···················· | ···················· | ···················· | ···················· | NNNNNNNNNNNNNNNNNNNN | NNNNNNNNNNNNNNNNNNNN | 1 |
| P | 6144 hCoV-19/Belgium/ULG-10581/2020\|EPI_ISL_626339\|2020 | ···················· | ···················· | ···················· | ···················· | NNNNNNNNNNNNNNNNNNNN | NNNNNNNNNNNNNNNNNNNN | 1 |
| P | 6146 hCoV-19/Cote dIvoire/BKE0201/2020\|EPI_ISL_614361\|2 | ···················· | ···················· | ···················· | ···················· | NNNNNNNNNNNNNNNNNNNN | NNNNNNNNNNNNNNNNNNNN | 1 |
| P | 6152 hCoV-19/Cote dIvoire/BKE0256/2020\|EPI_ISL_614367\|2 | ···················· | ···················· | ···················· | ···················· | NNNNNNNNNNNNNNNNNNNN | NNNNNNNNNNNNNNNNNNNN | 1 |
| P | 6155 hCoV-19/Cote dIvoire/BKE0317/2020\|EPI_ISL_614372\|2 | ···················· | ···················· | ···················· | ···················· | NNNNNNNNNNNNNNNNNNNN | NNNNNNNNNNNNNNNNNNNN | 1 |
| P | 6156 hCoV-19/Cote dIvoire/BKE0405/2020\|EPI_ISL_614375\|2 | ···················· | ···················· | ···················· | ···················· | NNNNNNNNNNNNNNNNNNNN | NNNNNNNNNNNNNNNNNNNN | 1 |
| P | 6160 hCoV-19/Cote dIvoire/BKE0633/2020\|EPI_ISL_614380\|2 | ···················· | ···················· | ···················· | ···················· | NNNNNNNNNNNNNNNNNNNN | NNNNNNNNNNNNNNNNNNNN | 1 |
| P | 6161 hCoV-19/Cote dIvoire/BKE0700/2020\|EPI_ISL_614381\|2 | ···················· | ···················· | ···················· | ···················· | NNNNNNNNNNNNNNNNNNNN | NNNNNNNNNNNNNNNNNNNN | 1 |
| P | 6164 hCoV-19/Cote dIvoire/BKE0880/2020\|EPI_ISL_614387\|2 | ···················· | ···················· | ···················· | ···················· | NNNNNNNNNNNNNNNNNNNN | NNNNNNNNNNNNNNNNNNNN | 1 |
| P | 6324 hCoV-19/New Zealand/20CV0310/2020\|EPI_ISL_622800\|2 | ···················· | ···················· | ···················· | ···················· | NNNNNNNNNNNNNNNNNNNN | NNNNNNNNNNNNNNNNNNNN | 1 |
| P | 6331 hCoV-19/New Zealand/20CV0413/2020\|EPI_ISL_622807\|2 | ···················· | ···················· | ···················· | ···················· | NNNNNNNNNNNNNNNNNNNN | NNNNNNNNNNNNNNNNNNNN | 1 |
| P | 6332 hCoV-19/New Zealand/20CV0426/2020\|EPI_ISL_622808\|2 | ···················· | ···················· | ···················· | ···················· | NNNNNNNNNNNNNNNNNNNN | NNNNNNNNNNNNNNNNNNNN | 1 |
| P | 6848 hCoV-19/Belgium/ULG-10624/2020\|EPI_ISL_626232\|2020 | ···················· | ···················· | ···················· | ···················· | NNNNNNNNNNNNNNNNNNNN | NNNNNNNNNNNNNNNNNNNN | 1 |
| P | 6849 hCoV-19/Belgium/ULG-10621/2020\|EPI_ISL_626233\|2020 | ···················· | ···················· | ·N·················· | ···················· | NNNNNNNNNNNNNNNNNNNN | NNNNNNNNNNNNNNNNNNNN | 1 |
| P | 6850 hCoV-19/Belgium/ULG-10625/2020\|EPI_ISL_626234\|2020 | ···················· | ···················· | ···················· | ···················· | NNNNNNNNNNNNNNNNNNNN | NNNNNNNNNNNNNNNNNNNN | 1 |
| P | 6855 hCoV-19/Belgium/ULG-10602/2020\|EPI_ISL_626239\|2020 | ···················· | ···················· | ···················· | ···················· | NNNNNNNNNNNNNNNNNNNN | NNNNNNNNNNNNNNNNNNNN | 1 |
| P | 6857 hCoV-19/Belgium/ULG-10618/2020\|EPI_ISL_626241\|2020 | ···················· | ···················· | ···················· | ···················· | NNNNNNNNNNNNNNNNNNNN | NNNNNNNNNNNNNNNNNNNN | 1 |
| P | 6858 hCoV-19/Belgium/ULG-10607/2020\|EPI_ISL_626242\|2020 | ···················· | ···················· | ···················· | ···················· | NNNNNNNNNNNNNNNNNNNN | NNNNNNNNNNNNNNNNNNNN | 1 |
| P | 6859 hCoV-19/Belgium/ULG-10616/2020\|EPI_ISL_626243\|2020 | ···················· | ···················· | ···················· | ···················· | NNNNNNNNNNNNNNNNNNNN | NNNNNNNNNNNNNNNNNNNN | 1 |
| P | 6860 hCoV-19/Belgium/ULG-10605/2020\|EPI_ISL_626244\|2020 | ···················· | ···················· | ···················· | ···················· | NNNNNNNNNNNNNNNNNNNN | NNNNNNNNNNNNNNNNNNNN | 1 |
| P | 6861 hCoV-19/Belgium/ULG-10610/2020\|EPI_ISL_626245\|2020 | ···················· | ···················· | ···················· | ···················· | NNNNNNNNNNNNNNNNNNNN | NNNNNNNNNNNNNNNNNNNN | 1 |
| P | 6868 hCoV-19/Belgium/ULG-10619/2020\|EPI_ISL_626252\|2020 | ···················· | ···················· | ···················· | ···················· | NNNNNNNNNNNNNNNNNNNN | NNNNNNNNNNNNNNNNNNNN | 1 |
| P | 6869 hCoV-19/Belgium/ULG-10613/2020\|EPI_ISL_626253\|2020 | ···················· | ·A·················· | ···················· | ···················· | NNNNNNNNNNNNNNNNNNNN | NNNNNNNNNNNNNNNNNNNN | 1 |
| P | 6870 hCoV-19/Belgium/ULG-10617/2020\|EPI_ISL_626254\|2020 | ···················· | ···················· | ···················· | ···················· | NNNNNNNNNNNNNNNNNNNN | NNNNNNNNNNNNNNNNNNNN | 1 |
| P | 6871 hCoV-19/Belgium/ULG-10580/2020\|EPI_ISL_626255\|2020 | ···················· | ···················· | ···················· | ···················· | NNNNNNNNNNNNNNNNNNNN | NNNNNNNNNNNNNNNNNNNN | 1 |
| P | 6873 hCoV-19/Belgium/ULG-10559/2020\|EPI_ISL_626257\|2020 | ···················· | ···················· | ···················· | ···················· | NNNNNNNNNNNNNNNNNNNN | NNNNNNNNNNNNNNNNNNNN | 1 |
| P | 6877 hCoV-19/Belgium/ULG-10597/2020\|EPI_ISL_626261\|2020 | ···················· | ···················· | ···················· | ···················· | NNNNNNNNNNNNNNNNNNNN | NNNNNNNNNNNNNNNNNNNN | 1 |
| P | 6878 hCoV-19/Belgium/ULG-10574/2020\|EPI_ISL_626262\|2020 | ···················· | ···················· | ···················· | ···················· | NNNNNNNNNNNNNNNNNNNN | NNNNNNNNNNNNNNNNNNNN | 1 |
| P | 6881 hCoV-19/Belgium/ULG-10542/2020\|EPI_ISL_626266\|2020 | ···················· | ···················· | ···················· | ···················· | NNNNNNNNNNNNNNNNNNNN | NNNNNNNNNNNNNNNNNNNN | 1 |
| P | 6882 hCoV-19/Belgium/ULG-10576/2020\|EPI_ISL_626267\|2020 | ···················· | ···················· | ···················· | ···················· | NNNNNNNNNNNNNNNNNNNN | NNNNNNNNNNNNNNNNNNNN | 1 |

| | | | | | | | | | |
|---|---|---|---|---|---|---|---|---|---|
| P | 6883 | hCoV-19/Belgium/ULG-10557/2020\|EPI_ISL_626268\|2020 | ···················· | ···················· | ···················· | ···················· | NNNNNNNNNNNNNNNNNNNN | NNNNNNNNNNNNNNNNNNNN | 1 |
| P | 6885 | hCoV-19/Belgium/ULG-10593/2020\|EPI_ISL_626270\|2020 | ···················· | ···················· | ···················· | ···················· | NNNNNNNNNNNNNNNNNNNN | NNNNNNNNNNNNNNNNNNNN | 1 |
| P | 6889 | hCoV-19/Belgium/ULG-10573/2020\|EPI_ISL_626274\|2020 | ···················· | ···················· | ···················· | ···················· | NNNNNNNNNNNNNNNNNNNN | NNNNNNNNNNNNNNNNNNNN | 1 |
| P | 6892 | hCoV-19/Belgium/ULG-10553/2020\|EPI_ISL_626277\|2020 | ···················· | ···················· | ···················· | ···················· | NNNNNNNNNNNNNNNNNNNN | NNNNNNNNNNNNNNNNNNNN | 1 |
| P | 6895 | hCoV-19/Belgium/ULG-10569/2020\|EPI_ISL_626280\|2020 | ···················· | ···················· | ···················· | ···················· | NNNNNNNNNNNNNNNNNNNN | NNNNNNNNNNNNNNNNNNNN | 1 |
| P | 6904 | hCoV-19/Belgium/ULG-10596/2020\|EPI_ISL_626289\|2020 | ···················· | ···················· | ···················· | ···················· | NNNNNNNNNNNNNNNNNNNN | NNNNNNNNNNNNNNNNNNNN | 1 |
| P | 6910 | hCoV-19/Belgium/ULG-10598/2020\|EPI_ISL_626296\|2020 | ···················· | ···················· | ···················· | ···················· | NNNNNNNNNNNNNNNNNNNN | NNNNNNNNNNNNNNNNNNNN | 1 |
| P | 7404 | hCoV-19/USA/CA-CZB-11958/2020\|EPI_ISL_625662\|2020- | ···················· | ···················· | ···················· | ···················· | ··NN················ | NNNNNNNNNNNNNNN····N | 1 |
| P | 7911 | hCoV-19/USA/CA-CZB-11932/2020\|EPI_ISL_625608\|2020- | ···················· | ·A·················· | ···················· | ···················· | NNNNNNNNNNNNNNNNNNNN | ···················· | 1 |
| P | 7920 | hCoV-19/USA/CA-CZB-11868/2020\|EPI_ISL_625621\|2020- | ···················· | ·A·················· | ···················· | ···················· | NNNNNNNNNNNNNNNNNNNN | NNNNNNNNNNNNNNNNNNNN | 1 |
| P | 7934 | hCoV-19/USA/CA-CZB-11802/2020\|EPI_ISL_625645\|2020- | ···················· | ·A·················· | ···················· | ···················· | NNNNNNNNNNNNNNNNNNNN | ···················· | 1 |
| P | 7935 | hCoV-19/USA/CA-CZB-11804/2020\|EPI_ISL_625647\|2020- | ···················· | ···················· | ···················· | ···················· | NNNNNNNNNNNNNNNNN··· | ···················· | 1 |
| P | 8124 | hCoV-19/USA/CA-CZB-11174/2020\|EPI_ISL_625530\|2020- | ···················· | ·A·················· | ···················· | ···················· | NNNNNNNNNNNNNNNNNNNN | ···················· | 1 |
| P | 8126 | hCoV-19/USA/CA-CZB-11176/2020\|EPI_ISL_625532\|2020- | ···················· | ·A·················· | ···················· | ···················· | NNNNNNNNNNNNNNNNNNNN | ···················· | 1 |
| P | 8129 | hCoV-19/USA/CA-CZB-11182/2020\|EPI_ISL_625536\|2020- | ···················· | ·A·················· | ···················· | ···················· | NNNNNNNNNNNNNNNNNNNN | NNNNNNNNNNNNNNNNNNNN | 1 |
| P | 8134 | hCoV-19/USA/CA-CZB-11187/2020\|EPI_ISL_625541\|2020- | ···················· | ·A·················· | ···················· | ···················· | NNNNNNNNNNNNNNNNNNNN | NNNNNNNNNNNNNNNNNNNN | 1 |
| P | 8136 | hCoV-19/USA/CA-CZB-11189/2020\|EPI_ISL_625543\|2020- | ···················· | ···················· | ···················· | ···················· | NNNNNNNNNNNNNN······ | ···················· | 1 |
| P | 8137 | hCoV-19/USA/CA-CZB-11190/2020\|EPI_ISL_625544\|2020- | ···················· | ·A·················· | ···················· | ···················· | NNNNNNNNNNNNNNNNNNNN | NNNNNNNNNNNNNNNNNNNN | 1 |
| P | 8139 | hCoV-19/USA/CA-CZB-11694/2020\|EPI_ISL_625546\|2020- | ···················· | ···················· | ···················· | ···················· | NNNNNNNNNNNNNNNNN··· | ···················· | 1 |
| P | 8156 | hCoV-19/USA/CA-CZB-11583/2020\|EPI_ISL_625565\|2020- | ···················· | ·A·················· | ···················· | ···················· | NNNNNNNNNNNNNNNNNNNN | NNNNNNNNNNNNNNNNNNNN | 1 |
| P | 8165 | hCoV-19/USA/CA-CZB-11523/2020\|EPI_ISL_625574\|2020- | ···················· | ···················· | ···················· | ···················· | NNNNNNNNNNNNNN······ | ···················· | 1 |
| P | 8166 | hCoV-19/USA/CA-CZB-11522/2020\|EPI_ISL_625575\|2020- | ···················· | ···················· | ···················· | ···················· | ···················N | NNNNNNNNNNNNNNNNNNNN | 1 |
| P | 8171 | hCoV-19/USA/CA-CZB-11516/2020\|EPI_ISL_625581\|2020- | ···················· | ·A·················· | ···················· | ···················· | NNNNNNNNNNNNNNNNNNNN | ···················· | 1 |
| P | 8192 | hCoV-19/USA/CA-CZB-11925/2020\|EPI_ISL_625603\|2020- | ···················· | ···················· | ···················· | ···················· | NNNNNNNNNNNNNNNNNNNN | ···················· | 1 |
| P | 8323 | hCoV-19/Denmark/DCGC-8488/2020\|EPI_ISL_618382\|2020 | ···················· | ···················· | ···················· | ···················· | NNNNNNNNNNNNNNNNNNNN | NNNNNNNNNNNNNNNNNNNN | 1 |
| P | 8445 | hCoV-19/USA/CA-CZB-11104/2020\|EPI_ISL_625500\|2020- | ···················· | ·R·················· | ···················· | ···················· | NNNNNNNNNNNNNNNNNNNN | NNNNNNNNNNNNNNNNNNNN | 1 |
| P | 8665 | hCoV-19/Bangladesh/CHRF-0047/2020\|EPI_ISL_625461\|2 | ···················· | ···················· | ···················· | ···················· | NNNNNNNNNNNNNNNNNNNN | NNNNNNNNNNNNNNNNNNNN | 1 |
| P | 8672 | hCoV-19/Bangladesh/CHRF-0063/2020\|EPI_ISL_625476\|2 | ···················· | ···················· | ···················· | ···················· | NNNNNNNNNNNNNNNNNNNN | ···················· | 1 |
| P | 8677 | hCoV-19/USA/CA-CZB-11652/2020\|EPI_ISL_625481\|2020- | ···················· | ·A·················· | ···················· | ···················· | NNNNNNNNNNNNNNNNNNNN | ···················· | 1 |
| P | 8687 | hCoV-19/USA/CA-CZB-11115/2020\|EPI_ISL_625508\|2020- | ···················· | ···················· | ···················· | ···················· | NNNNNNNNNNNNNNNNNNNN | NNNNNNNNNNNNNNNNNNNN | 1 |
| P | 8688 | hCoV-19/USA/CA-CZB-11116/2020\|EPI_ISL_625509\|2020- | ···················· | ···················· | ···················· | ···················· | NNNNNNNNNNNNNNNNNNNN | NNNNNNNNNNNNNNNNNNNN | 1 |
| P | 8689 | hCoV-19/USA/CA-CZB-11117/2020\|EPI_ISL_625510\|2020- | ···················· | ···················· | ···················· | ···················· | NNNNNNNNNNNNNNNNNNNN | NNNNNNNNNNNNNNNNNNNN | 1 |
| P | 8691 | hCoV-19/USA/CA-CZB-11119/2020\|EPI_ISL_625512\|2020- | ···················· | ···················· | ···················· | ···················· | NNNNNNNNNNNNNNNN···· | ···················· | 1 |
| P | 8693 | hCoV-19/USA/CA-CZB-11124/2020\|EPI_ISL_625515\|2020- | ···················· | ···················· | ···················· | ···················· | NNNNNNNNNNNNNNNNNNNN | ·NNN····NNNNNNN····· | 1 |
| P | 8695 | hCoV-19/USA/CA-CZB-11126/2020\|EPI_ISL_625517\|2020- | ···················· | ···················· | ···················· | ···················· | NNNNNNNNNNNNNNNNNNNN | NNNNNNNNNNNNNNN····· | 1 |
| P | 8696 | hCoV-19/USA/CA-CZB-11128/2020\|EPI_ISL_625518\|2020- | ···················· | ···················· | ···················· | ···················· | NNNNNNNNNNNNNNNNNNNN | NNNN···············N | 1 |
| P | 8698 | hCoV-19/USA/CA-CZB-11131/2020\|EPI_ISL_625520\|2020- | ···················· | ···················· | ···················· | ···················· | ····NNNN············ | ···················· | 1 |
| Q | 9 | hCoV-19/England/PORT-2D7246/2020\|EPI_ISL_628328\|20 | ···················· | ···················· | ···················· | ···················· | NNNNNNNNNNNNNNNNNNNN | NNNNNNNNNNNNNNNNNNNN | 1 |
| Q | 276 | hCoV-19/Wales/PHWC-485B95/2020\|EPI_ISL_628259\|2020 | ···················· | ···················· | ···················· | ···················· | NNNNNNNNNNNNNNNNNNNN | ···················· | 1 |
| Q | 281 | hCoV-19/Wales/PHWC-485CDE/2020\|EPI_ISL_628272\|2020 | ···················· | ···················· | ···················· | ···················· | NNNNNNNNNNNNNNNNNNNN | NNNNNNNNNNNNNNNNNNNN | 1 |
| Q | 289 | hCoV-19/Wales/PHWC-485DBD/2020\|EPI_ISL_628280\|2020 | ···················· | ···················· | ···················· | ···················· | ···NNNNNNNNNNNNNNNNN | ···················· | 1 |
| Q | 316 | hCoV-19/Wales/PHWC-4860A4/2020\|EPI_ISL_628307\|2020 | ···················· | ···················· | ···················· | ···················· | NNNNNNNNNNNNNNNNNNNN | NNNNNNNNNNNNNNNNNNNN | 1 |
| Q | 518 | hCoV-19/Wales/PHWC-485C38/2020\|EPI_ISL_628267\|2020 | ···················· | ···················· | ···················· | ···················· | NNNNNNNNNNNNNNNNNNNN | ···················· | 1 |
| Q | 696 | hCoV-19/Belgium/ULG-10688/2020\|EPI_ISL_636660\|2020 | ···················· | ···················· | ···················· | ···················· | NNNNNNNNNNNNNNNNNNNN | NNNNNNNNNNNNNNNNNNNN | 1 |
| Q | 697 | hCoV-19/Belgium/ULG-10689/2020\|EPI_ISL_636661\|2020 | ···················· | ···················· | ···················· | ···················· | NNNNNNNNNNNNNNNNNNNN | NNNNNNNNNNNNNNNNNNNN | 1 |
| Q | 698 | hCoV-19/Belgium/ULG-10690/2020\|EPI_ISL_636662\|2020 | ···················· | ···················· | ···················· | ···················· | NNNNNNNNNNNNNNNNNNNN | NNNNNNNNNNNNNNNNNNNN | 1 |
| Q | 699 | hCoV-19/Belgium/ULG-10691/2020\|EPI_ISL_636663\|2020 | ···················· | ···················· | ···················· | ···················· | NNNNNNNNNNNNNNNNNNNN | NNNNNNNNNNNNNNNNNNNN | 1 |
| Q | 700 | hCoV-19/Belgium/ULG-10692/2020\|EPI_ISL_636664\|2020 | ···················· | ···················· | ···················· | ···················· | NNNNNNNNNNNNNNNNNNNN | NNNNNNNNNNNNNNNNNNNN | 1 |
| Q | 701 | hCoV-19/Belgium/ULG-10693/2020\|EPI_ISL_636665\|2020 | ···················· | ···················· | ···················· | ···················· | NNNNNNNNNNNNNNNNNNNN | NNNNNNNNNNNNNNNNNNNN | 1 |
| Q | 702 | hCoV-19/Belgium/ULG-10694/2020\|EPI_ISL_636666\|2020 | ···················· | ···················· | ···················· | ···················· | NNNNNNNNNNNNNNNNNNNN | NNNNNNNNNNNNNNNNNNNN | 1 |
| Q | 703 | hCoV-19/Belgium/ULG-10695/2020\|EPI_ISL_636667\|2020 | ···················· | ···················· | ···················· | ···················· | NNNNNNNNNNNNNNNNNNNN | NNNNNNNNNNNNNNNNNNNN | 1 |
| Q | 704 | hCoV-19/Belgium/ULG-10696/2020\|EPI_ISL_636668\|2020 | ···················· | ···················· | ···················· | ···················· | NNNNNNNNNNNNNNNNNNNN | NNNNNNNNNNNNNNNNNNNN | 1 |
| Q | 705 | hCoV-19/Belgium/ULG-10697/2020\|EPI_ISL_636669\|2020 | ···················· | ···················· | ···················· | ···················· | NNNNNNNNNNNNNNNNNNNN | NNNNNNNNNNNNNNNNNNNN | 1 |
| Q | 706 | hCoV-19/Belgium/ULG-10698/2020\|EPI_ISL_636670\|2020 | ···················· | ···················· | ···················· | ···················· | NNNNNNNNNNNNNNNNNNNN | NNNNNNNNNNNNNNNNNNNN | 1 |
| Q | 707 | hCoV-19/Belgium/ULG-10699/2020\|EPI_ISL_636671\|2020 | ···················· | ···················· | ···················· | ···················· | NNNNNNNNNNNNNNNNNNNN | NNNNNNNNNNNNNNNNNNNN | 1 |
| Q | 709 | hCoV-19/Belgium/ULG-10701/2020\|EPI_ISL_636673\|2020 | ···················· | ···················· | ···················· | ···················· | NNNNNNNNNNNNNNNNNNNN | NNNNNNNNNNNNNNNNNNNN | 1 |
| Q | 710 | hCoV-19/Belgium/ULG-10702/2020\|EPI_ISL_636674\|2020 | ···················· | ···················· | ···················· | ···················· | NNNNNNNNNNNNNNNNNNNN | NNNNNNNNNNNNNNNNNNNN | 1 |
| Q | 712 | hCoV-19/Belgium/ULG-10704/2020\|EPI_ISL_636676\|2020 | ···················· | ···················· | ···················· | ···················· | NNNNNNNNNNNNNNNNNNNN | NNNNNNNNNNNNNNNNNNNN | 1 |
| Q | 713 | hCoV-19/Belgium/ULG-10705/2020\|EPI_ISL_636677\|2020 | ···················· | ···················· | ···················· | ···················· | NNNNNNNNNNNNNNNNNNNN | NNNNNNNNNNNNNNNNNNNN | 1 |
| Q | 714 | hCoV-19/Belgium/ULG-10707/2020\|EPI_ISL_636679\|2020 | ···················· | ···················· | ···················· | ···················· | NNNNNNNNNNNNNNNNNNNN | NNNNNNNNNNNNNNNNNNNN | 1 |
| Q | 774 | hCoV-19/Wales/PHWC-4856A3/2020\|EPI_ISL_628205\|2020 | ···················· | ···················· | ···················· | ···················· | NNNNNNNNNNNNNNNNNNNN | ···················· | 1 |
| Q | 780 | hCoV-19/Wales/PHWC-485728/2020\|EPI_ISL_628211\|2020 | ···················· | ···················· | ···················· | ···················· | ··NNNNNNNNNNNNNNNNNN | ···················· | 1 |
| Q | 1009 | hCoV-19/Belgium/ULG-10709/2020\|EPI_ISL_636680\|2020 | ···················· | ···················· | ···················· | ···················· | NNNNNNNNNNNNNNNNNNNN | NNNNNNNNNNNNNNNNNNNN | 1 |
| Q | 1010 | hCoV-19/Belgium/ULG-10710/2020\|EPI_ISL_636681\|2020 | ···················· | ···················· | ···················· | ···················· | NNNNNNNNNNNNNNNNNNNN | NNNNNNNNNNNNNNNNNNNN | 1 |
| Q | 1011 | hCoV-19/Belgium/ULG-10711/2020\|EPI_ISL_636682\|2020 | ···················· | ···················· | ···················· | ···················· | NNNNNNNNNNNNNNNNNNNN | NNNNNNNNNNNNNNNNNNNN | 1 |
| Q | 1012 | hCoV-19/Belgium/ULG-10713/2020\|EPI_ISL_636684\|2020 | ···················· | ···················· | ···················· | ···················· | NNNNNNNNNNNNNNNNNNNN | NNNNNNNNNNNNNNNNNNNN | 1 |
| Q | 1013 | hCoV-19/Belgium/ULG-10714/2020\|EPI_ISL_636685\|2020 | ···················· | ···················· | ···················· | ···················· | NNNNNNNNNNNNNNNNNNNN | NNNNNNNNNNNNNNNNNNNN | 1 |
| Q | 1028 | hCoV-19/Wales/PHWC-484E9D/2020\|EPI_ISL_628123\|2020 | ···················· | ···················· | ···················· | ···················· | NNNNNNNNNNNNNNNNNNNN | NNNNNNNNNNNNNNNNNNNN | 1 |
| Q | 1039 | hCoV-19/Wales/PHWC-484FD6/2020\|EPI_ISL_628134\|2020 | ···················· | ···················· | ···················· | ···················· | ··NNNNNNNNNNNN······ | ···················· | 1 |
| Q | 1068 | hCoV-19/Wales/PHWC-485290/2020\|EPI_ISL_628163\|2020 | ···················· | ···················· | ···················· | ···················· | NNNNNNNNNNNNNNNNNNNN | ···················· | 1 |
| Q | 1088 | hCoV-19/Wales/PHWC-485421/2020\|EPI_ISL_628183\|2020 | ···················· | ···················· | ···················· | ···················· | NNNNNNNNNNNNNNNNNNNN | ···················· | 1 |
| Q | 1249 | hCoV-19/Belgium/ULG-10638/2020\|EPI_ISL_636616\|2020 | ···················· | ···················· | ···················· | ···················· | NNNNNNNNNNNNNNNNNNNN | NNNNNNNNNNNNNNNNNNNN | 1 |
| Q | 1250 | hCoV-19/Belgium/ULG-10639/2020\|EPI_ISL_636617\|2020 | ···················· | ···················· | ···················· | ···················· | NNNNNNNNNNNNNNNNNNNN | NNNNNNNNNNNNNNNNNNNN | 1 |
| Q | 1251 | hCoV-19/Belgium/ULG-10641/2020\|EPI_ISL_636619\|2020 | ···················· | ···················· | ···················· | ···················· | NNNNNNNNNNNNNNNNNNNN | NNNNNNNNNNNNNNNNNNNN | 1 |
| Q | 1252 | hCoV-19/Belgium/ULG-10642/2020\|EPI_ISL_636620\|2020 | ···················· | ···················· | ···················· | ···················· | NNNNNNNNNNNNNNNNNNNN | NNNNNNNNNNNNNNNNNNNN | 1 |
| Q | 1253 | hCoV-19/Belgium/ULG-10643/2020\|EPI_ISL_636621\|2020 | ···················· | ···················· | ···················· | ···················· | NNNNNNNNNNNNNNNNNNNN | NNNNNNNNNNNNNNNNNNNN | 1 |
| Q | 1255 | hCoV-19/Belgium/ULG-10645/2020\|EPI_ISL_636623\|2020 | ···················· | ···················· | ···················· | ···················· | NNNNNNNNNNNNNNNNNNNN | NNNNNNNNNNNNNNNNNNNN | 1 |
| Q | 1256 | hCoV-19/Belgium/ULG-10646/2020\|EPI_ISL_636624\|2020 | ···················· | ···················· | ···················· | ···················· | NNNNNNNNNNNNNNNNNNNN | NNNNNNNNNNNNNNNNNNNN | 1 |
| Q | 1257 | hCoV-19/Belgium/ULG-10647/2020\|EPI_ISL_636625\|2020 | ···················· | ···················· | ···················· | ···················· | NNNNNNNNNNNNNNNNNNNN | NNNNNNNNNNNNNNNNNNNN | 1 |
| Q | 1258 | hCoV-19/Belgium/ULG-10648/2020\|EPI_ISL_636626\|2020 | ···················· | ···················· | ···················· | ···················· | NNNNNNNNNNNNNNNNNNNN | NNNNNNNNNNNNNNNNNNNN | 1 |

| | | | | | | | | | |
|---|---|---|---|---|---|---|---|---|---|
| Q | 1259 | hCoV-19/Belgium/ULG-10649/2020\|EPI_ISL_636627\|2020 | ···················· | ···················· | ···················· | ···················· | NNNNNNNNNNNNNNNNNNNN | NNNNNNNNNNNNNNNNNNNN | 1 |
| Q | 1260 | hCoV-19/Belgium/ULG-10650/2020\|EPI_ISL_636628\|2020 | ···················· | ···················· | ···················· | ···················· | NNNNNNNNNNNNNNNNNNNN | NNNNNNNNNNNNNNNNNNNN | 1 |
| Q | 1262 | hCoV-19/Belgium/ULG-10653/2020\|EPI_ISL_636630\|2020 | ···················· | ···················· | ···················· | ···················· | NNNNNNNNNNNNNNNNNNNN | NNNNNNNNNNNNNNNNNNNN | 1 |
| Q | 1267 | hCoV-19/Belgium/ULG-10658/2020\|EPI_ISL_636635\|2020 | ···················· | ···················· | ···················· | ···················· | NNNNNNNNNNNNNNNNNNNN | NNNNNNNNNNNNNNNNNNNN | 1 |
| Q | 1268 | hCoV-19/Belgium/ULG-10659/2020\|EPI_ISL_636636\|2020 | ···················· | ···················· | ···················· | ···················· | NNNNNNNNNNNNNNNNNNNN | NNNNNNNNNNNNNNNNNNNN | 1 |
| Q | 1270 | hCoV-19/Belgium/ULG-10675/2020\|EPI_ISL_636649\|2020 | ···················· | ···················· | ···················· | ···················· | NNNNNNNNNNNNNNNNNNNN | NNNNNNNNNNNNNNNNNNNN | 1 |
| Q | 1271 | hCoV-19/Belgium/ULG-10677/2020\|EPI_ISL_636650\|2020 | ···················· | ···················· | ···················· | ···················· | NNNNNNNNNNNNNNNNNNNN | NNNNNNNNNNNNNNNNNNNN | 1 |
| Q | 1272 | hCoV-19/Belgium/ULG-10678/2020\|EPI_ISL_636651\|2020 | ···················· | ···················· | ···················· | ···················· | NNNNNNNNNNNNNNNNNNNN | NNNNNNNNNNNNNNNNNNNN | 1 |
| Q | 1273 | hCoV-19/Belgium/ULG-10680/2020\|EPI_ISL_636652\|2020 | ···················· | ···················· | ···················· | ···················· | NNNNNNNNNNNNNNNNNNNN | NNNNNNNNNNNNNNNNNNNN | 1 |
| Q | 1274 | hCoV-19/Belgium/ULG-10681/2020\|EPI_ISL_636653\|2020 | ···················· | ···················· | ···················· | ···················· | NNNNNNNNNNNNNNNNNNNN | NNNNNNNNNNNNNNNNNNNN | 1 |
| Q | 1275 | hCoV-19/Belgium/ULG-10682/2020\|EPI_ISL_636654\|2020 | ···················· | ···················· | ···················· | ···················· | NNNNNNNNNNNNNNNNNNNN | NNNNNNNNNNNNNNNNNNNN | 1 |
| Q | 1276 | hCoV-19/Belgium/ULG-10683/2020\|EPI_ISL_636655\|2020 | ···················· | ···················· | ···················· | ···················· | NNNNNNNNNNNNNNNNNNNN | NNNNNNNNNNNNNNNNNNNN | 1 |
| Q | 1278 | hCoV-19/Belgium/ULG-10685/2020\|EPI_ISL_636657\|2020 | ···················· | ···················· | ···················· | ···················· | NNNNNNNNNNNNNNNNNNNN | NNNNNNNNNNNNNNNNNNNN | 1 |
| Q | 1280 | hCoV-19/Belgium/ULG-10687/2020\|EPI_ISL_636659\|2020 | ···················· | ·A·················· | ···················· | ···················· | NNNNNNNNNNNNNNNNNNNN | NNNNNNNNNNNNNNNNNNNN | 1 |
| Q | 1372 | hCoV-19/Slovenia/5333/2020\|EPI_ISL_635217\|2020-03- | ···················· | ···················· | ···················· | ···················· | NNNNNNNNNNNNNNNNNNNN | NNNNNNNNNNNNNNNNNNNN | 1 |
| Q | 1406 | hCoV-19/Slovenia/10734/2020\|EPI_ISL_635253\|2020-03 | ···················· | ···················· | ···················· | ···················· | NNNNNNNNNNNNNNNNNNNN | NNNNNNNNNNNNNNNNNNNN | 1 |
| Q | 1408 | hCoV-19/Slovenia/10986/2020\|EPI_ISL_635255\|2020-03 | ···················· | ···················· | ···················· | ···················· | ·NNNNNNNNNNNNNNNNNNN | ···················· | 1 |
| Q | 1410 | hCoV-19/Slovenia/11976/2020\|EPI_ISL_635257\|2020-03 | ···················· | ···················· | ···················· | ···················· | NNNNNNNNNNNNNNNNNNNN | NNNNNNNNNNNNNNNNNNNN | 1 |
| Q | 1412 | hCoV-19/Slovenia/12380/2020\|EPI_ISL_635259\|2020-03 | ···················· | ···················· | ···················· | ···················· | NNNNNNNNNNNNNNNNNNNN | NNNNNNNNNNNNNNNNNNNN | 1 |
| Q | 1416 | hCoV-19/Slovenia/13022/2020\|EPI_ISL_635263\|2020-03 | ···················· | ···················· | ···················· | ···················· | NNNNNNNNNNNNNNNNNNNN | NNNNNNNNNNNNNNNNNNNN | 1 |
| Q | 1423 | hCoV-19/Slovenia/V0-14718/2020\|EPI_ISL_635271\|2020 | ···················· | NNNNNNNNNNNNNNNNNNNN | ···················· | ···················· | ···················· | ···················· | 1 |
| Q | 1515 | hCoV-19/Belgium/ULG-10626/2020\|EPI_ISL_636605\|2020 | ···················· | ···················· | ···················· | ···················· | NNNNNNNNNNNNNNNNNNNN | NNNNNNNNNNNNNNNNNNNN | 1 |
| Q | 1516 | hCoV-19/Belgium/ULG-10627/2020\|EPI_ISL_636606\|2020 | ···················· | ···················· | ···················· | ···················· | NNNNNNNNNNNNNNNNNNNN | NNNNNNNNNNNNNNNNNNNN | 1 |
| Q | 1517 | hCoV-19/Belgium/ULG-10628/2020\|EPI_ISL_636607\|2020 | ···················· | ···················· | ···················· | ···················· | NNNNNNNNNNNNNNNNNNNN | NNNNNNNNNNNNNNNNNNNN | 1 |
| Q | 1518 | hCoV-19/Belgium/ULG-10629/2020\|EPI_ISL_636608\|2020 | ···················· | ···················· | ···················· | ···················· | NNNNNNNNNNNNNNNNNNNN | NNNNNNNNNNNNNNNNNNNN | 1 |
| Q | 1519 | hCoV-19/Belgium/ULG-10630/2020\|EPI_ISL_636609\|2020 | ···················· | ···················· | ···················· | ···················· | NNNNNNNNNNNNNNNNNNNN | NNNNNNNNNNNNNNNNNNNN | 1 |
| Q | 1521 | hCoV-19/Belgium/ULG-10632/2020\|EPI_ISL_636611\|2020 | ···················· | ···················· | ···················· | ···················· | NNNNNNNNNNNNNNNNNNNN | NNNNNNNNNNNNNNNNNNNN | 1 |
| Q | 1524 | hCoV-19/Belgium/ULG-10636/2020\|EPI_ISL_636614\|2020 | ···················· | ···················· | ···················· | ···················· | NNNNNNNNNNNNNNNNNNNN | NNNNNNNNNNNNNNNNNNNN | 1 |
| Q | 1528 | hCoV-19/Belgium/ULG-10663/2020\|EPI_ISL_636640\|2020 | ···················· | ···················· | ···················· | ···················· | NNNNNNNNNNNNNNNNNNNN | NNNNNNNNNNNNNNNNNNNN | 1 |
| Q | 1532 | hCoV-19/Belgium/ULG-10668/2020\|EPI_ISL_636644\|2020 | ···················· | ···················· | ···················· | ···················· | NNNNNNNNNNNNNNNNNNNN | NNNNNNNNNNNNNNNNNNNN | 1 |
| Q | 1533 | hCoV-19/Belgium/ULG-10669/2020\|EPI_ISL_636645\|2020 | ···················· | ···················· | ···················· | ···················· | NNNNNNNNNNNNNNNNNNNN | NNNNNNNNNNNNNNNNNNNN | 1 |
| Q | 1534 | hCoV-19/Belgium/ULG-10671/2020\|EPI_ISL_636646\|2020 | ···················· | ···················· | ···················· | ···················· | NNNNNNNNNNNNNNNNNNNN | NNNNNNNNNNNNNNNNNNNN | 1 |
| Q | 1539 | hCoV-19/Wales/PHWC-48449B/2020\|EPI_ISL_628008\|2020 | ···················· | ···················· | ···················· | ···················· | NNNNNNNNNNNNNNNNNNNN | NNNNNNNNNNNNNNNNNNNN | 1 |
| Q | 1546 | hCoV-19/Wales/PHWC-48455C/2020\|EPI_ISL_628015\|2020 | ···················· | ···················· | ···················· | ···················· | ·····NNNNNNNNNNNNNNN | ···················· | 1 |
| Q | 1547 | hCoV-19/Wales/PHWC-484589/2020\|EPI_ISL_628016\|2020 | ···················· | ···················· | ···················· | ···················· | NNNNNNNNNNNNNNNNNNNN | ···················· | 1 |
| Q | 1551 | hCoV-19/Wales/PHWC-484756/2020\|EPI_ISL_628040\|2020 | ···················· | ···················· | ···················· | ···················· | ·······NNNNNNNNNNNNN | NNNNNNNNNNNNNNN····· | 1 |
| Q | 1572 | hCoV-19/Wales/PHWC-484941/2020\|EPI_ISL_628061\|2020 | ···················· | ···················· | ···················· | ···················· | ···················· | NNNNNNNNNN·········· | 1 |
| Q | 1587 | hCoV-19/Wales/PHWC-484AB7/2020\|EPI_ISL_628076\|2020 | ···················· | ···················· | ···················· | ···················· | NNNNNNNNNNNNNNNNNNNN | ···················· | 1 |
| Q | 1593 | hCoV-19/Wales/PHWC-484D27/2020\|EPI_ISL_628102\|2020 | ···················· | ···················· | ···················· | ···················· | NNNNNNNNNNNN········ | ···················· | 1 |
| Q | 1789 | hCoV-19/Slovenia/V0-6255/2020\|EPI_ISL_635295\|2020- | ···················· | NNNNNNNNNNNNNNNNNNNN | ···················· | ···················· | ···················· | ···················· | 1 |
| Q | 1794 | hCoV-19/Wales/PHWC-4843F8/2020\|EPI_ISL_628001\|2020 | ···················· | ···················· | ···················· | ···················· | NNNNNNNNNNNNNNNNNNNN | NNNNNNNNNNNNNNNNNNNN | 1 |
| Q | 1795 | hCoV-19/Wales/PHWC-484404/2020\|EPI_ISL_628002\|2020 | ···················· | ···················· | ···················· | ···················· | NNNNNNNNNNNNNNNNNNNN | NNNNNNNNNNNNNNNNNNNN | 1 |
| Q | 1796 | hCoV-19/Wales/PHWC-484413/2020\|EPI_ISL_628003\|2020 | ···················· | ···················· | ···················· | ···················· | NNNNNNNNNNNNNNNNNNNN | ···················· | 1 |
| Q | 1805 | hCoV-19/Wales/PHWC-48464A/2020\|EPI_ISL_628026\|2020 | ···················· | ···················· | ···················· | ···················· | NNNNNNNNNNNNNNN····· | ···················· | 1 |
| Q | 1813 | hCoV-19/Wales/PHWC-4846D1/2020\|EPI_ISL_628034\|2020 | ···················· | ···················· | ···················· | ···················· | NNNN················ | ···················· | 1 |
| Q | 1827 | hCoV-19/Wales/PHWC-484BE1/2020\|EPI_ISL_628088\|2020 | ···················· | ···················· | ···················· | ···················· | ··NNNNNNNNNNNNNNNNNN | NNNNNNNNNNNNNNNNNNNN | 1 |
| Q | 1831 | hCoV-19/Wales/PHWC-484C57/2020\|EPI_ISL_628092\|2020 | ···················· | ···················· | ···················· | ···················· | ·NNNNNNN··NN········ | ···················· | 1 |
| Q | 1836 | hCoV-19/Wales/PHWC-484CC0/2020\|EPI_ISL_628097\|2020 | ···················· | ···················· | ···················· | ···················· | NNNNNNNNNNNNNNNNNNNN | NNNNNNNNNNNNNNNNN··· | 1 |
| Q | 2542 | hCoV-19/USA/WI-UW-1882/2020\|EPI_ISL_632897\|2020-09 | ···················· | ···················· | ···················· | ···················· | NNNNNNNNNNNNNNNNNNNN | NNNNNNNNNNNNNNNNNNNN | 1 |
| Q | 2568 | hCoV-19/Scotland/CVR5225/2020\|EPI_ISL_627397\|2020- | ···················· | ···················· | ···················· | ···················· | NN············NN··NN | NN·N···NN··········· | 1 |
| Q | 2570 | hCoV-19/Scotland/CVR5228/2020\|EPI_ISL_627399\|2020- | ···················· | ···················· | ···················· | ···················· | NNNNNNNNNNNNNNNNNNNN | NNNNNNNNNNNNNNNNNNNN | 1 |
| Q | 2576 | hCoV-19/England/EXET-1388E8/2020\|EPI_ISL_627430\|20 | ···················· | ···················· | ···················· | ···················· | NNNNNNNNNNNNNNNNNNNN | NNNNNNNNNNNNNNNNNNNN | 1 |
| Q | 2590 | hCoV-19/England/EXET-138DCB/2020\|EPI_ISL_627444\|20 | ···················· | ·A·················· | ···················· | ···················· | NNNNNNNNNNNNNNNNNNNN | NNNNNNNNNNNNNNNNNNNN | 1 |
| Q | 2633 | hCoV-19/England/MILK-A19125/2020\|EPI_ISL_627487\|20 | ···················· | ···················· | ···················· | ···················· | ·····NNNNNNNNNNNNNNN | ···················· | 1 |
| Q | 2817 | hCoV-19/England/NORW-F10D5/2020\|EPI_ISL_627300\|202 | ···················· | ···················· | ···················· | ···················· | NNNNNNNNNNNNNNNNNNNN | NNNNNNNNNNNNNNNNNNNN | 1 |
| Q | 3073 | hCoV-19/England/NOTT-1196D4/2020\|EPI_ISL_627310\|20 | ···················· | ···················· | ···················· | ···················· | NNNNNNNNNNNNNNNNNNNN | NNNNNNNNNNNNNNNNNNNN | 1 |
| Q | 3077 | hCoV-19/England/CAMC-A66332/2020\|EPI_ISL_627317\|20 | ···················· | ·A·······A·········· | ···················· | ···················· | NNNNNNNNNNNNNNNNNNNN | NNNNNNNNNNNNNNNNNNNN | 1 |
| Q | 3082 | hCoV-19/England/CAMC-A666A2/2020\|EPI_ISL_627322\|20 | ···················· | ···················· | ···················· | ···················· | NNNNNNNNNNNNNNNNNNNN | NNNNNNNNNNNNNNNNNNNN | 1 |
| Q | 3140 | hCoV-19/Scotland/CVR5182/2020\|EPI_ISL_627381\|2020- | ···················· | ···················· | ···················· | ···················· | ······N·······N····· | NN···········NN····· | 1 |
| Q | 3148 | hCoV-19/Scotland/CVR5207/2020\|EPI_ISL_627389\|2020- | ···················· | ···················· | ···················· | ···················· | ···················· | NN·····NN··········· | 1 |
| Q | 3330 | hCoV-19/Wales/PHWC-482FAB/2020\|EPI_ISL_627295\|2020 | ···················· | ···················· | ···················· | ···················· | NNNNNNNNNNNNNNNNNNNN | NNNNNNNNNNNNNNNNNNNN | 1 |
| Q | 3586 | hCoV-19/Wales/PHWC-484B0F/2020\|EPI_ISL_627204\|2020 | ···················· | ···················· | ···················· | ···················· | ···················· | NNNNNNNNNNNNNNN····· | 1 |
| Q | 3603 | hCoV-19/Wales/PHWC-485931/2020\|EPI_ISL_627242\|2020 | ···················· | ···················· | ···················· | ···················· | ···················· | NNNNNNNNNNNNNNNNNNNN | 1 |
| Q | 3857 | hCoV-19/Wales/PHWC-483C67/2020\|EPI_ISL_627225\|2020 | ···················· | ···················· | ···················· | ···················· | NNNNNNNNNNNNNNNNNNNN | NNNNNNNNNNNNNNNNNNNN | 1 |
| Q | 3860 | hCoV-19/Wales/PHWC-484282/2020\|EPI_ISL_627228\|2020 | ···················· | ···················· | ···················· | ···················· | NNNNNNNNNNNNNNNNNNNN | ···················· | 1 |
| Q | 4066 | hCoV-19/Canada/ON-PHL-20-01756/2020\|EPI_ISL_636909 | ···················· | ······A············· | ···················· | ···················· | NNNNNNNNNNNNNNNNNNNN | ···················· | 1 |
| Q | 4067 | hCoV-19/Canada/ON-PHL-20-01757/2020\|EPI_ISL_636910 | ···················· | ······A············· | ···················· | ···················· | NNNNN··············· | ···················· | 1 |
| Q | 4076 | hCoV-19/Canada/ON-PHL-20-01766/2020\|EPI_ISL_636919 | ···················· | ···················· | ···················· | ···················· | NNNNNNNNNNNNNNNNNNNN | NNNNNNNNNNNNNNNNNNNN | 1 |
| Q | 4078 | hCoV-19/Canada/ON-PHL-20-01768/2020\|EPI_ISL_636921 | ···················· | ···················· | ···················· | ···················· | NNNNNNNNNNNNNNNNNNNN | NNNNNNNNNNNNNNNNNNNN | 1 |
| Q | 4082 | hCoV-19/Canada/ON-PHL-20-01772/2020\|EPI_ISL_636925 | ···················· | ······A············· | ···················· | ···················· | NNNNNNNNNNNNNNNNNNNN | NNNNNNNNNNNNNNNNNNNN | 1 |
| Q | 4083 | hCoV-19/Canada/ON-PHL-20-01775/2020\|EPI_ISL_636927 | ···················· | ···················· | ···················· | ···················· | NNNNNNNNNNNNNNNNNNNN | ···················· | 1 |
| Q | 4085 | hCoV-19/Canada/ON-PHL-20-01777/2020\|EPI_ISL_636929 | ···················· | ···················· | ···················· | ···················· | NNNNNNNNNNNNNNNNNNNN | NNNNNNNNNNNNNNNNNNNN | 1 |
| Q | 4088 | hCoV-19/Canada/ON-PHL-20-01781/2020\|EPI_ISL_636932 | ···················· | ···················· | ···················· | ···················· | NNNNNNNNNNNNN······· | ···················· | 1 |
| Q | 4089 | hCoV-19/Canada/ON-PHL-20-01782/2020\|EPI_ISL_636933 | ···················· | ···················· | ···················· | ···················· | NNNNNNNNNNNNNNNNNNNN | NNNNNNNNNNNNNNNNNNNN | 1 |
| Q | 4319 | hCoV-19/Canada/ON-PHL-20-01785/2020\|EPI_ISL_636935 | ···················· | ···················· | ···················· | ···················· | NNNN················ | ···················· | 1 |
| Q | 4323 | hCoV-19/Canada/ON-PHL-20-01789/2020\|EPI_ISL_636939 | ···················· | ······A············· | ···················· | ···················· | NNNNNNNNNNNNNNNNNNNN | NNNNNNNNNNNNNNNNNNNN | 1 |
| Q | 4326 | hCoV-19/Canada/ON-PHL-20-01792/2020\|EPI_ISL_636942 | ···················· | ···················· | ···················· | ···················· | NNNNNNNNNNNNNNNNNNNN | ···················· | 1 |
| Q | 4327 | hCoV-19/Canada/ON-PHL-20-01793/2020\|EPI_ISL_636943 | ···················· | ···················· | ···················· | ···················· | NNNNNNNNNNNNNNNNNNNN | NNNNNNNNNNNNNNNNNNNN | 1 |
| Q | 4329 | hCoV-19/Canada/ON-PHL-20-01795/2020\|EPI_ISL_636945 | ···················· | ···················· | ···················· | ···················· | NNNN················ | ···················· | 1 |
| Q | 4332 | hCoV-19/Canada/ON-PHL-20-01805/2020\|EPI_ISL_636949 | ···················· | ···················· | ···················· | ···················· | NNNN················ | ···················· | 1 |

| | | | | | | | | |
|---|---|---|---|---|---|---|---|---|
| Q | 4337 hCoV-19/Canada/ON-PHL-20-01810/2020\|EPI_ISL_636954 | ···················· | ···················· | ···················· | ···················· | NNNNNN·············· | ··········NNNNNNNNNN | 1 |
| Q | 4339 hCoV-19/Canada/ON-PHL-20-01812/2020\|EPI_ISL_636956 | ···················· | ···················· | ···················· | ···················· | NNNNNNNNNNNNNNNNN··· | NNNNNNNNNNNNNNNNNNNN | 1 |
| Q | 4355 hCoV-19/England/NORW-F2238/2020\|EPI_ISL_627072\|202 | ···················· | ···················· | ···················· | ···················· | NNNNNNNNNNNNNNNNNNNN | NNNNNNNNNNNNNNN··NNN | 1 |
| Q | 4357 hCoV-19/England/NORW-F32FB/2020\|EPI_ISL_627074\|202 | ···················· | ···················· | ···················· | ···················· | NNNNNNNNNNNNNNNNNNNN | NNNNNNNNNNNNNNNNNNNN | 1 |
| Q | 4358 hCoV-19/England/NORW-F35F2/2020\|EPI_ISL_627075\|202 | ···················· | ···················· | ···················· | ···················· | NNNNNNNNNNNNNNNNNNNN | NNNNN··············· | 1 |
| Q | 4652 hCoV-19/Wales/PHWC-48365A/2020\|EPI_ISL_627841\|2020 | ···················· | ···················· | ···················· | ···················· | NNNNNNNNNNNN········ | NNNNNNNNNNNNNNNNNNNN | 1 |
| Q | 4654 hCoV-19/Wales/PHWC-483678/2020\|EPI_ISL_627843\|2020 | ···················· | ···················· | ···················· | ···················· | ·NNNNNNNNNNNNNNNNNNN | NNNNNNNNNNNNNNNNNNNN | 1 |
| Q | 4655 hCoV-19/Wales/PHWC-483687/2020\|EPI_ISL_627844\|2020 | ···················· | ···················· | ···················· | ···················· | NNNNNNNNNNNNNNNNNNNN | ···················· | 1 |
| Q | 4659 hCoV-19/Wales/PHWC-4836D2/2020\|EPI_ISL_627848\|2020 | ···················· | ···················· | ···················· | ···················· | ··NNNNNNNNNNNNNNNNNN | NNNNNNNNNNNNNNNNNNNN | 1 |
| Q | 4663 hCoV-19/Wales/PHWC-483924/2020\|EPI_ISL_627873\|2020 | ···················· | ···················· | ···················· | ···················· | NNNNNNNNNNNNNNNNNNNN | NNNNNNNNNNNNNNNNNNNN | 1 |
| Q | 4668 hCoV-19/Wales/PHWC-48398E/2020\|EPI_ISL_627878\|2020 | ···················· | ···················· | ···················· | ···················· | NNNNNNNNNNNNNNNNNNNN | NNNNNNNNNNNNNNN····· | 1 |
| Q | 4670 hCoV-19/Wales/PHWC-4839BB/2020\|EPI_ISL_627880\|2020 | ···················· | ···················· | ···················· | ···················· | ···················· | NNNNNNNN············ | 1 |
| Q | 4676 hCoV-19/Wales/PHWC-483A4F/2020\|EPI_ISL_627886\|2020 | ···················· | ···················· | ···················· | ···················· | NNNNNNNNNNNNNN······ | ···················· | 1 |
| Q | 4680 hCoV-19/Wales/PHWC-483AA9/2020\|EPI_ISL_627890\|2020 | ···················· | ···················· | ···················· | ···················· | ·NNNNNNNNNNNNNNNN··· | ···················· | 1 |
| Q | 4911 hCoV-19/Wales/PHWC-4833AE/2020\|EPI_ISL_627811\|2020 | ···················· | ···················· | ···················· | ···················· | ·NNNNNNNNNNNNNNNNNNN | ···················· | 1 |
| Q | 4924 hCoV-19/Wales/PHWC-4834AB/2020\|EPI_ISL_627824\|2020 | ···················· | ···················· | ···················· | ···················· | NNNNNNNNNNNNNNNNNNNN | ···················· | 1 |
| Q | 4929 hCoV-19/Wales/PHWC-483599/2020\|EPI_ISL_627834\|2020 | ···················· | ···················· | ···················· | ···················· | ·NNNNNNNNNNNNNNNNNNN | ···················· | 1 |
| Q | 4943 hCoV-19/Wales/PHWC-4837EE/2020\|EPI_ISL_627859\|2020 | ···················· | ···················· | ···················· | ···················· | NNNNNNNNNNNNNNNNNNNN | ···················· | 1 |
| Q | 4947 hCoV-19/Wales/PHWC-483863/2020\|EPI_ISL_627863\|2020 | ···················· | ···················· | ···················· | ···················· | ··NNNNNNNNNNNNNNN··· | ···················· | 1 |
| Q | 4949 hCoV-19/Wales/PHWC-483881/2020\|EPI_ISL_627865\|2020 | ···················· | ···················· | ···················· | ···················· | ··NN·········NNNN··· | ·······NNN·········· | 1 |
| Q | 4952 hCoV-19/Wales/PHWC-4838CD/2020\|EPI_ISL_627868\|2020 | ···················· | ···················· | ···················· | ···················· | ··NNNNNNNNNNNNNNNNNN | ···················· | 1 |
| Q | 5106 hCoV-19/South Africa/KRISP-K003991/2020\|EPI_ISL_63 | ···················· | ···················· | ···················· | ···················· | NNNNNNNNNNNNNNNNNNNN | NNNNNNNNNNNNNNNNNNNN | 1 |
| Q | 5204 hCoV-19/Wales/PHWC-482D65/2020\|EPI_ISL_627735\|2020 | ···················· | ···················· | ···················· | ···················· | NNNNNNNNNNNNNNNNNNNN | ···················· | 1 |
| Q | 5207 hCoV-19/Wales/PHWC-482DA1/2020\|EPI_ISL_627738\|2020 | ···················· | ···················· | ···················· | ···················· | ·NNNNNNNNNNNNNNNNNNN | ···················· | 1 |
| Q | 5213 hCoV-19/Wales/PHWC-482E08/2020\|EPI_ISL_627744\|2020 | ···················· | ···················· | ···················· | ···················· | ····NNNNNNNNNNNNNNNN | NNNNNNNNN··········· | 1 |
| Q | 5216 hCoV-19/Wales/PHWC-482E35/2020\|EPI_ISL_627747\|2020 | ···················· | ···················· | ···················· | ···················· | ···················· | NNNNNNNNNNNNNNN····· | 1 |
| Q | 5217 hCoV-19/Wales/PHWC-482E44/2020\|EPI_ISL_627748\|2020 | ···················· | ···················· | ···················· | ···················· | NNNNNNNNNNNN········ | ···················· | 1 |
| Q | 5218 hCoV-19/Wales/PHWC-482E62/2020\|EPI_ISL_627749\|2020 | ···················· | ···················· | ···················· | ···················· | ·NNNNNNNNNNNNNNNNNNN | NNNN················ | 1 |
| Q | 5219 hCoV-19/Wales/PHWC-482EBD/2020\|EPI_ISL_627750\|2020 | ···················· | ···················· | ···················· | ···················· | ··NNNNNNNNNN········ | ···················· | 1 |
| Q | 5220 hCoV-19/Wales/PHWC-482ECC/2020\|EPI_ISL_627751\|2020 | ···················· | ···················· | ···················· | ···················· | ···················· | NNNN················ | 1 |
| Q | 5250 hCoV-19/Wales/PHWC-4830F2/2020\|EPI_ISL_627781\|2020 | ···················· | ···················· | ···················· | ···················· | NNNNNNNNNNNNNNNNNNNN | ···················· | 1 |
| Q | 5263 hCoV-19/Wales/PHWC-483256/2020\|EPI_ISL_627794\|2020 | ···················· | ···················· | ···················· | ···················· | ·NNNNNNNNNNNNNNNNNNN | NNNNNNNNNNNNNNNNNNNN | 1 |
| Q | 5268 hCoV-19/Wales/PHWC-4832DE/2020\|EPI_ISL_627799\|2020 | ···················· | ···················· | ···················· | ···················· | NNNNNNNNNNNNNNNNNNNN | NNNNN··············· | 1 |
| Q | 5583 hCoV-19/France/ARA-SC186/2020\|EPI_ISL_634836\|2020- | ···················· | ···················· | ···················· | NNNNNNNNNNNNNNNNNNNN | ···················· | ···················· | 1 |
| Q | 5613 hCoV-19/Belgium/UGent-155/2020\|EPI_ISL_634878\|2020 | ···················· | ···················· | ···················· | ···················· | NNNNNNNNNNNNNNNNNNNN | NNNNNNNNNNNNNNNNNNNN | 1 |
| Q | 5615 hCoV-19/Belgium/UGent-161/2020\|EPI_ISL_634880\|2020 | ···················· | ···················· | ···················· | ···················· | NNNNNNNNNNNNNNNNNNNN | NNNNNNNNNNNNNNNNNNNN | 1 |
| Q | 5621 hCoV-19/Belgium/UGent-170/2020\|EPI_ISL_634886\|2020 | ···················· | ···················· | ···················· | ···················· | NNNNNNNNNNNNNNNNNNNN | NNNNNNNNNNNNNNNNNNNN | 1 |
| Q | 5625 hCoV-19/Belgium/UGent-179/2020\|EPI_ISL_634890\|2020 | ···················· | ···················· | ···················· | ···················· | NNNNNNNNNNNNNNNNNNNN | NNNNNNNNNNNNNNNNNNNN | 1 |
| Q | 5674 hCoV-19/England/NORW-F1512/2020\|EPI_ISL_627601\|202 | ···················· | ···················· | ···················· | ···················· | NNNNNNNNNNNNNNNNNNN· | ···················· | 1 |
| Q | 5677 hCoV-19/England/NORW-F156D/2020\|EPI_ISL_627604\|202 | ···················· | ···················· | ···················· | ···················· | NNNN···NNNNN········ | N··················· | 1 |
| Q | 5681 hCoV-19/England/NORW-F161F/2020\|EPI_ISL_627608\|202 | ···················· | ···················· | ···················· | ···················· | NNN················· | NNNN················ | 1 |
| Q | 5684 hCoV-19/England/NORW-F1679/2020\|EPI_ISL_627611\|202 | ···················· | ···················· | ···················· | ···················· | NNNNNNNNNNNNNNNNNNNN | NNNNNNNNNNNNNNNNNNNN | 1 |
| Q | 5686 hCoV-19/England/NORW-F16F1/2020\|EPI_ISL_627613\|202 | ···················· | ···················· | ···················· | ···················· | NNNNN··············· | ···················· | 1 |
| Q | 5688 hCoV-19/England/NORW-F17C1/2020\|EPI_ISL_627615\|202 | ···················· | ···················· | ···················· | ···················· | NNNNNNNNNNNNNNNNNNNN | NNNNNNNNNNNNNNNNNNNN | 1 |
| Q | 5689 hCoV-19/England/NORW-F17D0/2020\|EPI_ISL_627616\|202 | ···················· | ···················· | ···················· | ···················· | NNNNNNNNNNNNNNNNNNNN | NNNNNNNNNNNNNNNNNNNN | 1 |
| Q | 5703 hCoV-19/England/NORW-F1CD1/2020\|EPI_ISL_627630\|202 | ···················· | ···················· | ···················· | ···················· | NNNNNNNNNNNNNNNNNNNN | NNNNNNNNNNNNNNNNNNNN | 1 |
| Q | 5707 hCoV-19/England/NORW-F1D56/2020\|EPI_ISL_627634\|202 | ···················· | ···················· | ···················· | ···················· | ···················· | NNNNNNNNNNNNNNNN·N·N | 1 |
| Q | 5712 hCoV-19/England/NORW-F1E17/2020\|EPI_ISL_627639\|202 | ···················· | ···················· | ···················· | ···················· | ···················· | NNNNN··············· | 1 |
| Q | 5715 hCoV-19/England/NORW-F1E44/2020\|EPI_ISL_627642\|202 | ···················· | ···················· | ···················· | ···················· | NNNN················ | ···················· | 1 |
| Q | 5717 hCoV-19/England/NORW-F1F7E/2020\|EPI_ISL_627644\|202 | ···················· | ···················· | ···················· | ···················· | NNNN··NNN··········· | ···················· | 1 |
| Q | 5719 hCoV-19/England/NORW-F1FBA/2020\|EPI_ISL_627646\|202 | ···················· | ···················· | ···················· | ···················· | NNNN·NN············· | NNNNNN·············· | 1 |
| Q | 5732 hCoV-19/England/NORW-F2195/2020\|EPI_ISL_627659\|202 | ···················· | ···················· | ···················· | ···················· | NNNNNNNNNNNNNNNNNNNN | NNNNNNNNNNNNNNNNNNNN | 1 |
| Q | 5734 hCoV-19/England/NORW-F2317/2020\|EPI_ISL_627661\|202 | ···················· | ···················· | ···················· | ···················· | ···················· | NNNNNNNNNNNNNNNNNNNN | 1 |
| Q | 5735 hCoV-19/England/NORW-F239F/2020\|EPI_ISL_627662\|202 | ···················· | ···················· | ···················· | ···················· | NNN················· | NNNNN··············· | 1 |
| Q | 5846 hCoV-19/England/QEUH-AE4D98/2020\|EPI_ISL_633402\|20 | ···················· | NNNNNNNNNNNNNNNNNNNN | ···················· | ···················· | ···················· | ···················· | 1 |
| Q | 5919 hCoV-19/USA/MI-UM-10036456046/2020\|EPI_ISL_628847\| | ···················· | ···················· | ···················· | ···················· | NNNNNNNNNNNNNNNNNNNN | ···················· | 1 |
| Q | 5920 hCoV-19/USA/MI-UM-10036455010/2020\|EPI_ISL_628848\| | ···················· | ···················· | ···················· | ···················· | NNNNNNNNNNNNNNNNNNNN | ···················· | 1 |
| Q | 5924 hCoV-19/USA/MI-UM-10036495367/2020\|EPI_ISL_628856\| | ···················· | ···················· | ···················· | ···················· | NNNNNNNNNNNNNNNNNNNN | NNNNNNNNNNNNNNNNNNNN | 1 |
| Q | 5927 hCoV-19/USA/MI-UM-10036524330/2020\|EPI_ISL_628878\| | ···················· | ······A············· | ···················· | ···················· | NNNNNNNNNNNNNNNNNNNN | ···················· | 1 |
| Q | 6165 hCoV-19/Northern Ireland/NIRE-108B9D/2020\|EPI_ISL_ | ···················· | ···················· | ···················· | ···················· | ·····NNNNNNNNNNNNNNN | ···················· | 1 |
| Q | 6167 hCoV-19/Northern Ireland/NIRE-108BE8/2020\|EPI_ISL_ | ···················· | ···················· | ···················· | ···················· | ·NNNNNN············· | ···················· | 1 |
| Q | 6175 hCoV-19/Northern Ireland/NIRE-108CD6/2020\|EPI_ISL_ | ···················· | ···················· | ···················· | ···················· | NNNNNNNNNNNNNNNNNNNN | ···················· | 1 |
| Q | 6189 hCoV-19/Northern Ireland/NIRE-108E49/2020\|EPI_ISL_ | ···················· | ···················· | ···················· | ···················· | ···················· | NNNNNNNNNNNNNNN····· | 1 |
| Q | 6195 hCoV-19/England/NORW-F02A3/2020\|EPI_ISL_627572\|202 | ···················· | ···················· | ···················· | ···················· | NNNNNNNNNNNNNN····NN | ···················· | 1 |
| Q | 6200 hCoV-19/England/NORW-F02FE/2020\|EPI_ISL_627577\|202 | ···················· | ···················· | ···················· | ···················· | NNNNNNNNNNNNNNNNNNN· | NNNNN··············· | 1 |
| Q | 6201 hCoV-19/England/NORW-F0319/2020\|EPI_ISL_627578\|202 | ···················· | ···················· | ···················· | ···················· | NNNNNNNNNNNNNNNNNNNN | NNNNNNNNNNNNNNNNNNNN | 1 |
| Q | 6202 hCoV-19/England/NORW-F0337/2020\|EPI_ISL_627579\|202 | ···················· | ···················· | ···················· | ···················· | NNNNNNNNNNNNNNNNNNNN | NNNNNNNNNNNNNNNNNNNN | 1 |
| Q | 6205 hCoV-19/England/NORW-F0364/2020\|EPI_ISL_627582\|202 | ···················· | ···················· | ···················· | ···················· | ···················· | NNNNNNNNNNNNNNNN···· | 1 |
| Q | 6208 hCoV-19/England/NORW-F065C/2020\|EPI_ISL_627585\|202 | ···················· | ···················· | ···················· | ···················· | NNNNNNNNNNNNNNNNNNNN | NNNNNNNNNNNNNNNNNNNN | 1 |
| Q | 6209 hCoV-19/England/NORW-F0ECD/2020\|EPI_ISL_627586\|202 | ···················· | ···················· | ···················· | ···················· | NNNNNNNNNNNNNNNNNNNN | ···················· | 1 |
| Q | 6213 hCoV-19/England/NORW-F0F7F/2020\|EPI_ISL_627590\|202 | ···················· | ···················· | ···················· | ···················· | NNNNNNNNNNNNNNNNNNNN | NNNNNNNNNNNNNNNNNNNN | 1 |
| Q | 6215 hCoV-19/England/NORW-F0FBB/2020\|EPI_ISL_627592\|202 | ···················· | ···················· | ···················· | ···················· | NNNNNNNNNNNNNNNNNNNN | NNNNNNNNNNNNNNNNNNNN | 1 |
| Q | 6218 hCoV-19/England/NORW-F1178/2020\|EPI_ISL_627595\|202 | ···················· | ···················· | ···················· | ···················· | NNNNNNNNNNNNNNNNNNNN | NNNNNNNNNNNNNNNN·NNN | 1 |
| Q | 6229 hCoV-19/USA/MI-UM-10036460497/2020\|EPI_ISL_628810\| | ···················· | ···················· | ···················· | ···················· | NNNNNNNNNNNNNNNNNNNN | NNNNNNNNNNNNNNNNNNNN | 1 |
| Q | 6233 hCoV-19/USA/MI-UM-10036482555/2020\|EPI_ISL_628815\| | ···················· | ···················· | ···················· | ···················· | NNNNNNNNNNNNNNNNNNNN | ···················· | 1 |
| Q | 6234 hCoV-19/USA/MI-UM-10036454962/2020\|EPI_ISL_628816\| | ···················· | ···················· | ···················· | ···················· | NNNNNNNNNNNNNNNNNNNN | ···················· | 1 |
| Q | 6236 hCoV-19/USA/MI-UM-10036464855/2020\|EPI_ISL_628818\| | ···················· | ···················· | ···················· | ···················· | NNNNNNNNNNNNNNNNNNNN | NNNNNNNNNNNNNNNNNNNN | 1 |
| Q | 6241 hCoV-19/USA/MI-UM-10036463292/2020\|EPI_ISL_628823\| | ···················· | ···················· | ···················· | ···················· | NNNNNNNNNNNNNNNNNNNN | ···················· | 1 |
| Q | 6246 hCoV-19/USA/MI-UM-10036455024/2020\|EPI_ISL_628830\| | ···················· | ···················· | ···················· | ···················· | NNNNNNNNNNNNNNNNNNNN | NNNNNNNNNNNNNNNNNNNN | 1 |
| Q | 6253 hCoV-19/USA/MI-UM-10036501790/2020\|EPI_ISL_628860\| | ···················· | ···················· | ···················· | ···················· | NNNNNNNNNNNNNNNNNNNN | NNNNNNNNNNNNNNNNNNNN | 1 |

| | | | | | | | | |
|---|---|---|---|---|---|---|---|---|
| Q | 6255 hCoV-19/USA/MI-UM-10036501769/2020\|EPI_ISL_628862\| | ···················· | ···················· | ···················· | ···················· | NNNNNNNNNNNNNNNNNNNN | ···················· | 1 |
| Q | 6256 hCoV-19/USA/MI-UM-10036490275/2020\|EPI_ISL_628863\| | ···················· | ···················· | ···················· | ···················· | NNNNNNNNNNNNNNNNNNNN | NNNNNNNNNNNNNNNNNNNN | 1 |
| Q | 6257 hCoV-19/USA/MI-UM-10036524225/2020\|EPI_ISL_628867\| | ···················· | ···················· | ···················· | ···················· | NNNNNNNNNNNNNNNNNNNN | NNNNNNNNNNNNNNNNNNNN | 1 |
| Q | 6259 hCoV-19/USA/MI-UM-10036516572/2020\|EPI_ISL_628870\| | ···················· | ···················· | ···················· | ···················· | NNNNNNNNNNNNNNNNNNNN | ···················· | 1 |
| Q | 6260 hCoV-19/USA/MI-UM-10036507339/2020\|EPI_ISL_628873\| | ···················· | ···················· | ···················· | ···················· | NNNNNNNNNNNNNNNNNNNN | NNNNNNNNNNNNNNNNNNNN | 1 |
| Q | 6266 hCoV-19/USA/MI-UM-10036507377/2020\|EPI_ISL_628881\| | ···················· | ···················· | ···················· | ···················· | NNNNNNNNNNNNNNNNNNNN | NNNNNNNNNNNNNNNNNNNN | 1 |
| Q | 6268 hCoV-19/USA/MI-UM-10036522216/2020\|EPI_ISL_628884\| | ···················· | ···················· | ···················· | ···················· | NNNNNNNNNNNNNNNNNNNN | NNNNNNNNNNNNNNNNNNNN | 1 |
| Q | 6677 hCoV-19/USA/WI-WSLH-200703/2020\|EPI_ISL_631460\|202 | ···················· | ···················· | ···················· | ···················· | NNNNNNNNNNNNNNNNNNNN | NNNNNNNNNNNNNNNNNNNN | 1 |
| Q | 6678 hCoV-19/USA/WI-WSLH-200704/2020\|EPI_ISL_631461\|202 | ···················· | ···················· | ···················· | ···················· | NNNNNNNNNNNNNNNNNNNN | NNNNNNNNNNNNNNNNNNNN | 1 |
| Q | 6683 hCoV-19/USA/WI-WSLH-200709/2020\|EPI_ISL_631466\|202 | ···················· | ···················· | ···················· | ···················· | NNNNNNNNNNNNNNNNNNNN | NNNNNNNNNNNNNNNNNNNN | 1 |
| Q | 6684 hCoV-19/USA/WI-WSLH-200710/2020\|EPI_ISL_631467\|202 | ···················· | ···················· | ···················· | ···················· | NNNNNNNNNNNNNNNNNNNN | NNNNNNNNNNNNNNNNNNNN | 1 |
| Q | 6685 hCoV-19/USA/WI-WSLH-200745/2020\|EPI_ISL_631500\|202 | ···················· | ···················· | ···················· | ···················· | NNNNNNNNNNNNNNNNNNNN | NNNNNNNNNNNNNNNNNNNN | 1 |
| Q | 6686 hCoV-19/USA/WI-WSLH-200746/2020\|EPI_ISL_631501\|202 | ···················· | ···················· | ···················· | ···················· | NNNNNNNNNNNNNNNNNNNN | NNNNNNNNNNNNNNNNNNNN | 1 |
| Q | 6789 hCoV-19/South Africa/KRISP-K004068/2020\|EPI_ISL_63 | ···················· | NNNNNNNNNNNNNNNNNNNN | ···················· | ···················· | ···················· | ···················· | 1 |
| Q | 6916 hCoV-19/France/ARA-SC068/2020\|EPI_ISL_629104\|2020- | ···················· | ···················· | NNNNNNNN············ | NNNNNNNNNNNNNNNNNNNN | ···················· | ···················· | 1 |
| Q | 7035 hCoV-19/USA/WI-WSLH-200632/2020\|EPI_ISL_631402\|202 | ···················· | ···················· | ···················· | ···················· | NNNNNNNNNNNNNNNNNNNN | NNNNNNNNNNNNNNNNNNNN | 1 |
| Q | 7036 hCoV-19/USA/WI-WSLH-200635/2020\|EPI_ISL_631404\|202 | ···················· | ···················· | ···················· | ···················· | NNNNNNNNNNNNNNNNNNNN | NNNNNNNNNNNNNNNNNNNN | 1 |
| Q | 7037 hCoV-19/USA/WI-WSLH-200637/2020\|EPI_ISL_631406\|202 | ···················· | ···················· | ···················· | ···················· | NNNNNNNNNNNNNNNNNNNN | NNNNNNNNNNNNNNNNNNNN | 1 |
| Q | 7038 hCoV-19/USA/WI-WSLH-200638/2020\|EPI_ISL_631407\|202 | ·T·················· | ···················· | ···················· | ···················· | NNNNNNNNNNNNNNNNNNNN | NNNNNNNNNNNNNNNNNNNN | 1 |
| Q | 7039 hCoV-19/USA/WI-WSLH-200639/2020\|EPI_ISL_631408\|202 | ·T·················· | ···················· | ···················· | ···················· | NNNNNNNNNNNNNNNNNNNN | NNNNNNNNNNNNNNNNNNNN | 1 |
| Q | 7040 hCoV-19/USA/WI-WSLH-200640/2020\|EPI_ISL_631409\|202 | ···················· | ···················· | ···················· | ···················· | NNNNNNNNNNNNNNNNNNNN | NNNNNNNNNNNNNNNNNNNN | 1 |
| Q | 7041 hCoV-19/USA/WI-WSLH-200641/2020\|EPI_ISL_631410\|202 | ···················· | ···················· | ···················· | ···················· | NNNNNNNNNNNNNNNNNNNN | NNNNNNNNNNNNNNNNNNNN | 1 |
| Q | 7042 hCoV-19/USA/WI-WSLH-200643/2020\|EPI_ISL_631412\|202 | ···················· | ···················· | ···················· | ···················· | NNNNNNNNNNNNNNNNNNNN | NNNNNNNNNNNNNNNNNNNN | 1 |
| Q | 7043 hCoV-19/USA/WI-WSLH-200644/2020\|EPI_ISL_631413\|202 | ···················· | ···················· | ···················· | ···················· | NNNNNNNNNNNNNNNNNNNN | NNNNNNNNNNNNNNNNNNNN | 1 |
| Q | 7044 hCoV-19/USA/WI-WSLH-200645/2020\|EPI_ISL_631414\|202 | ···················· | ···················· | ···················· | ···················· | NNNNNNNNNNNNNNNNNNNN | NNNNNNNNNNNNNNNNNNNN | 1 |
| Q | 7045 hCoV-19/USA/WI-WSLH-200646/2020\|EPI_ISL_631415\|202 | ···················· | ···················· | ···················· | ···················· | NNNNNNNNNNNNNNNNNNNN | NNNNNNNNNNNNNNNNNNNN | 1 |
| Q | 7046 hCoV-19/USA/WI-WSLH-200647/2020\|EPI_ISL_631416\|202 | ···················· | ···················· | ···················· | ···················· | NNNNNNNNNNNNNNNNNNNN | NNNNNNNNNNNNNNNNNNNN | 1 |
| Q | 7047 hCoV-19/USA/WI-WSLH-200648/2020\|EPI_ISL_631417\|202 | ···················· | ···················· | ···················· | ···················· | NNNNNNNNNNNNNNNNNNNN | NNNNNNNNNNNNNNNNNNNN | 1 |
| Q | 7048 hCoV-19/USA/WI-WSLH-200651/2020\|EPI_ISL_631418\|202 | ···················· | ···················· | ···················· | ···················· | NNNNNNNNNNNNNNNNNNNN | NNNNNNNNNNNNNNNNNNNN | 1 |
| Q | 7049 hCoV-19/USA/WI-WSLH-200652/2020\|EPI_ISL_631419\|202 | ···················· | ···················· | ···················· | ···················· | NNNNNNNNNNNNNNNNNNNN | NNNNNNNNNNNNNNNNNNNN | 1 |
| Q | 7050 hCoV-19/USA/WI-WSLH-200653/2020\|EPI_ISL_631420\|202 | ···················· | ···················· | ···················· | ···················· | NNNNNNNNNNNNNNNNNNNN | NNNNNNNNNNNNNNNNNNNN | 1 |
| Q | 7051 hCoV-19/USA/WI-WSLH-200654/2020\|EPI_ISL_631421\|202 | ···················· | ···················· | ···················· | ···················· | NNNNNNNNNNNNNNNNNNNN | NNNNNNNNNNNNNNNNNNNN | 1 |
| Q | 7052 hCoV-19/USA/WI-WSLH-200655/2020\|EPI_ISL_631422\|202 | ···················· | ···················· | ···················· | ···················· | NNNNNNNNNNNNNNNNNNNN | NNNNNNNNNNNNNNNNNNNN | 1 |
| Q | 7053 hCoV-19/USA/WI-WSLH-200657/2020\|EPI_ISL_631423\|202 | ···················· | ···················· | ···················· | ···················· | NNNNNNNNNNNNNNNNNNNN | NNNNNNNNNNNNNNNNNNNN | 1 |
| Q | 7054 hCoV-19/USA/WI-WSLH-200658/2020\|EPI_ISL_631424\|202 | ···················· | ···················· | ···················· | ···················· | NNNNNNNNNNNNNNNNNNNN | NNNNNNNNNNNNNNNNNNNN | 1 |
| Q | 7057 hCoV-19/USA/WI-WSLH-200661/2020\|EPI_ISL_631427\|202 | ···················· | ···················· | ···················· | ···················· | NNNNNNNNNNNNNNNNNNNN | NNNNNNNNNNNNNNNNNNNN | 1 |
| Q | 7058 hCoV-19/USA/WI-WSLH-200663/2020\|EPI_ISL_631428\|202 | ···················· | ···················· | ···················· | ···················· | NNNNNNNNNNNNNNNNNNNN | NNNNNNNNNNNNNNNNNNNN | 1 |
| Q | 7059 hCoV-19/USA/WI-WSLH-200664/2020\|EPI_ISL_631429\|202 | ···················· | ···················· | ···················· | ···················· | NNNNNNNNNNNNNNNNNNNN | NNNNNNNNNNNNNNNNNNNN | 1 |
| Q | 7060 hCoV-19/USA/WI-WSLH-200665/2020\|EPI_ISL_631430\|202 | ···················· | ···················· | ···················· | ···················· | NNNNNNNNNNNNNNNNNNNN | NNNNNNNNNNNNNNNNNNNN | 1 |
| Q | 7061 hCoV-19/USA/WI-WSLH-200670/2020\|EPI_ISL_631431\|202 | ···················· | ···················· | ···················· | ···················· | NNNNNNNNNNNNNNNNNNNN | NNNNNNNNNNNNNNNNNNNN | 1 |
| Q | 7062 hCoV-19/USA/WI-WSLH-200674/2020\|EPI_ISL_631432\|202 | ···················· | ···················· | ···················· | ···················· | NNNNNNNNNNNNNNNNNNNN | NNNNNNNNNNNNNNNNNNNN | 1 |
| Q | 7064 hCoV-19/USA/WI-WSLH-200677/2020\|EPI_ISL_631434\|202 | ···················· | ···················· | ···················· | ···················· | NNNNNNNNNNNNNNNNNNNN | NNNNNNNNNNNNNNNNNNNN | 1 |
| Q | 7065 hCoV-19/USA/WI-WSLH-200678/2020\|EPI_ISL_631435\|202 | ···················· | ···················· | ···················· | ···················· | NNNNNNNNNNNNNNNNNNNN | NNNNNNNNNNNNNNNNNNNN | 1 |
| Q | 7066 hCoV-19/USA/WI-WSLH-200679/2020\|EPI_ISL_631436\|202 | ···················· | ···················· | ···················· | ···················· | NNNNNNNNNNNNNNNNNNNN | ···················· | 1 |
| Q | 7067 hCoV-19/USA/WI-WSLH-200680/2020\|EPI_ISL_631437\|202 | ···················· | ···················· | ···················· | ···················· | NNNNNNNNNNNNNNNNNNNN | NNNNNNNNNNNNNNNNNNNN | 1 |
| Q | 7068 hCoV-19/USA/WI-WSLH-200681/2020\|EPI_ISL_631438\|202 | ···················· | ···················· | ···················· | ···················· | NNNNNNNNNNNNNNNNNNNN | NNNNNNNNNNNNNNNNNNNN | 1 |
| Q | 7069 hCoV-19/USA/WI-WSLH-200682/2020\|EPI_ISL_631439\|202 | ···················· | ···················· | ···················· | ···················· | NNNNNNNNNNNNNNNNNNNN | NNNNNNNNNNNNNNNNNNNN | 1 |
| Q | 7070 hCoV-19/USA/WI-WSLH-200683/2020\|EPI_ISL_631440\|202 | ···················· | ···················· | ···················· | ···················· | NNNNNNNNNNNNNNNNNNNN | NNNNNNNNNNNNNNNNNNNN | 1 |
| Q | 7071 hCoV-19/USA/WI-WSLH-200684/2020\|EPI_ISL_631441\|202 | ···················· | ···················· | ···················· | ···················· | NNNNNNNNNNNNNNNNNNNN | NNNNNNNNNNNNNNNNNNNN | 1 |
| Q | 7072 hCoV-19/USA/WI-WSLH-200685/2020\|EPI_ISL_631442\|202 | ···················· | ···················· | ···················· | ···················· | NNNNNNNNNNNNNNNNNNNN | NNNNNNNNNNNNNNNNNNNN | 1 |
| Q | 7073 hCoV-19/USA/WI-WSLH-200686/2020\|EPI_ISL_631443\|202 | ···················· | ···················· | ···················· | ···················· | NNNNNNNNNNNNNNNNNNNN | NNNNNNNNNNNNNNNNNNNN | 1 |
| Q | 7074 hCoV-19/USA/WI-WSLH-200687/2020\|EPI_ISL_631444\|202 | ···················· | ···················· | ···················· | ···················· | NNNNNNNNNNNNNNNNNNNN | NNNNNNNNNNNNNNNNNNNN | 1 |
| Q | 7075 hCoV-19/USA/WI-WSLH-200688/2020\|EPI_ISL_631445\|202 | ···················· | ···················· | ···················· | ···················· | NNNNNNNNNNNNNNNNNNNN | NNNNNNNNNNNNNNNNNNNN | 1 |
| Q | 7076 hCoV-19/USA/WI-WSLH-200689/2020\|EPI_ISL_631446\|202 | ···················· | ···················· | ···················· | ···················· | NNNNNNNNNNNNNNNNNNNN | NNNNNNNNNNNNNNNNNNNN | 1 |
| Q | 7077 hCoV-19/USA/WI-WSLH-200690/2020\|EPI_ISL_631447\|202 | ···················· | ···················· | ···················· | ···················· | NNNNNNNNNNNNNNNNNNNN | NNNNNNNNNNNNNNNNNNNN | 1 |
| Q | 7079 hCoV-19/USA/WI-WSLH-200692/2020\|EPI_ISL_631449\|202 | ···················· | ···················· | ···················· | ···················· | NNNNNNNNNNNNNNNNNNNN | NNNNNNNNNNNNNNNNNNNN | 1 |
| Q | 7080 hCoV-19/USA/WI-WSLH-200693/2020\|EPI_ISL_631450\|202 | ···················· | ···················· | ···················· | ···················· | NNNNNNNNNNNNNNNNNNNN | NNNNNNNNNNNNNNNNNNNN | 1 |
| Q | 7081 hCoV-19/USA/WI-WSLH-200694/2020\|EPI_ISL_631451\|202 | ···················· | ···················· | ···················· | ···················· | NNNNNNNNNNNNNNNNNNNN | NNNNNNNNNNNNNNNNNNNN | 1 |
| Q | 7082 hCoV-19/USA/WI-WSLH-200695/2020\|EPI_ISL_631452\|202 | ···················· | ···················· | ···················· | ···················· | NNNNNNNNNNNNNNNNNNNN | NNNNNNNNNNNNNNNNNNNN | 1 |
| Q | 7083 hCoV-19/USA/WI-WSLH-200696/2020\|EPI_ISL_631453\|202 | ···················· | ···················· | ···················· | ···················· | NNNNNNNNNNNNNNNNNNNN | NNNNNNNNNNNNNNNNNNNN | 1 |
| Q | 7085 hCoV-19/USA/WI-WSLH-200698/2020\|EPI_ISL_631455\|202 | ···················· | ···················· | ···················· | ···················· | NNNNNNNNNNNNNNNNNNNN | NNNNNNNNNNNNNNNNNNNN | 1 |
| Q | 7086 hCoV-19/USA/WI-WSLH-200699/2020\|EPI_ISL_631456\|202 | ···················· | ···················· | ···················· | ···················· | NNNNNNNNNNNNNNNNNNNN | NNNNNNNNNNNNNNNNNNNN | 1 |
| Q | 7087 hCoV-19/USA/WI-WSLH-200700/2020\|EPI_ISL_631457\|202 | ···················· | ···················· | ···················· | ···················· | NNNNNNNNNNNNNNNNNNNN | NNNNNNNNNNNNNNNNNNNN | 1 |
| Q | 7088 hCoV-19/USA/WI-WSLH-200702/2020\|EPI_ISL_631459\|202 | ···················· | ···················· | ···················· | ···················· | NNNNNNNNNNNNNNNNNNNN | NNNNNNNNNNNNNNNNNNNN | 1 |
| Q | 7089 hCoV-19/USA/WI-WSLH-200711/2020\|EPI_ISL_631468\|202 | ···················· | ···················· | ···················· | ···················· | NNNNNNNNNNNNNNNNNNNN | NNNNNNNNNNNNNNNNNNNN | 1 |
| Q | 7090 hCoV-19/USA/WI-WSLH-200712/2020\|EPI_ISL_631469\|202 | ···················· | ···················· | ···················· | ···················· | NNNNNNNNNNNNNNNNNNNN | NNNNNNNNNNNNNNNNNNNN | 1 |
| Q | 7091 hCoV-19/USA/WI-WSLH-200713/2020\|EPI_ISL_631470\|202 | ···················· | ···················· | ···················· | ···················· | NNNNNNNNNNNNNNNNNNNN | NNNNNNNNNNNNNNNNNNNN | 1 |
| Q | 7092 hCoV-19/USA/WI-WSLH-200715/2020\|EPI_ISL_631471\|202 | ···················· | ···················· | ···················· | ···················· | NNNNNNNNNNNNNNNNNNNN | NNNNNNNNNNNNNNNNNNNN | 1 |
| Q | 7093 hCoV-19/USA/WI-WSLH-200716/2020\|EPI_ISL_631472\|202 | ···················· | ···················· | ···················· | ···················· | NNNNNNNNNNNNNNNNNNNN | NNNNNNNNNNNNNNNNNNNN | 1 |
| Q | 7094 hCoV-19/USA/WI-WSLH-200717/2020\|EPI_ISL_631473\|202 | ···················· | ···················· | ···················· | ···················· | NNNNNNNNNNNNNNNNNNNN | NNNNNNNNNNNNNNNNNNNN | 1 |
| Q | 7095 hCoV-19/USA/WI-WSLH-200718/2020\|EPI_ISL_631474\|202 | ···················· | ···················· | ···················· | ···················· | NNNNNNNNNNNNNNNNNNNN | NNNNNNNNNNNNNNNNNNNN | 1 |
| Q | 7096 hCoV-19/USA/WI-WSLH-200719/2020\|EPI_ISL_631475\|202 | ···················· | ···················· | ···················· | ···················· | NNNNNNNNNNNNNNNNNNNN | NNNNNNNNNNNNNNNNNNNN | 1 |
| Q | 7097 hCoV-19/USA/WI-WSLH-200720/2020\|EPI_ISL_631476\|202 | ···················· | ···················· | ···················· | ···················· | NNNNNNNNNNNNNNNNNNNN | NNNNNNNNNNNNNNNNNNNN | 1 |
| Q | 7098 hCoV-19/USA/WI-WSLH-200721/2020\|EPI_ISL_631477\|202 | ···················· | ·A·················· | ···················· | ···················· | NNNNNNNNNNNNNNNNNNNN | NNNNNNNNNNNNNNNNNNNN | 1 |
| Q | 7103 hCoV-19/USA/WI-WSLH-200726/2020\|EPI_ISL_631482\|202 | ···················· | ···················· | ···················· | ···················· | NNNNNNNNNNNNNNNNNNNN | NNNNNNNNNNNNNNNNNNNN | 1 |
| Q | 7108 hCoV-19/USA/WI-WSLH-200731/2020\|EPI_ISL_631487\|202 | ···················· | ···················· | ···················· | ···················· | NNNNNNNNNNNNNNNNNNNN | NNNNNNNNNNNNNNNNNNNN | 1 |
| Q | 7109 hCoV-19/USA/WI-WSLH-200732/2020\|EPI_ISL_631488\|202 | ···················· | ···················· | ···················· | ···················· | NNNNNNNNNNNNNNNNNNNN | NNNNNNNNNNNNNNNNNNNN | 1 |
| Q | 7110 hCoV-19/USA/WI-WSLH-200733/2020\|EPI_ISL_631489\|202 | ···················· | ···················· | ···················· | ···················· | NNNNNNNNNNNNNNNNNNNN | NNNNNNNNNNNNNNNNNNNN | 1 |
| Q | 7250 hCoV-19/USA/WI-WSLH-200607/2020\|EPI_ISL_631400\|202 | ···················· | ···················· | ···················· | ···················· | NNNNNNNNNNNNNNNNNNNN | NNNNNNNNNNNNNNNNNNNN | 1 |
| Q | 7251 hCoV-19/USA/WI-WSLH-200608/2020\|EPI_ISL_631401\|202 | ···················· | ···················· | ···················· | ···················· | NNNNNNNNNNNNNNNNNNNN | NNNNNNNNNNNNNNNNNNNN | 1 |

| | | | | | | | | | |
|---|---|---|---|---|---|---|---|---|---|
| Q | 7252 | hCoV-19/USA/WI-WSLH-200734/2020\|EPI_ISL_631490\|202 | ···················· | ···················· | ···················· | ···················· | NNNNNNNNNNNNNNNNNNNN | NNNNNNNNNNNNNNNNNNNN | 1 |
| Q | 7253 | hCoV-19/USA/WI-WSLH-200735/2020\|EPI_ISL_631491\|202 | ···················· | ···················· | ···················· | ···················· | NNNNNNNNNNNNNNNNNNNN | NNNNNNNNNNNNNNNNNNNN | 1 |
| Q | 7254 | hCoV-19/USA/WI-WSLH-200736/2020\|EPI_ISL_631492\|202 | ···················· | ···················· | ···················· | ···················· | NNNNNNNNNNNNNNNNNNNN | NNNNNNNNNNNNNNNNNNNN | 1 |
| Q | 7255 | hCoV-19/USA/WI-WSLH-200737/2020\|EPI_ISL_631493\|202 | ···················· | ···················· | ···················· | ···················· | NNNNNNNNNNNNNNNNNNNN | NNNNNNNNNNNNNNNNNNNN | 1 |
| Q | 7256 | hCoV-19/USA/WI-WSLH-200738/2020\|EPI_ISL_631494\|202 | ···················· | ···················· | ···················· | ···················· | NNNNNNNNNNNNNNNNNNNN | NNNNNNNNNNNNNNNNNNNN | 1 |
| Q | 7258 | hCoV-19/USA/WI-WSLH-200741/2020\|EPI_ISL_631496\|202 | ···················· | ···················· | ···················· | ···················· | NNNNNNNNNNNNNNNNNNNN | NNNNNNNNNNNNNNNNNNNN | 1 |
| Q | 7259 | hCoV-19/USA/WI-WSLH-200742/2020\|EPI_ISL_631497\|202 | ···················· | ···················· | ···················· | ···················· | NNNNNNNNNNNNNNNNNNNN | NNNNNNNNNNNNNNNNNNNN | 1 |
| Q | 7261 | hCoV-19/USA/WI-WSLH-200744/2020\|EPI_ISL_631499\|202 | ···················· | ···················· | ···················· | ···················· | NNNNNNNNNNNNNNNNNNNN | NNNNNNNNNNNNNNNNNNNN | 1 |
| Q | 7572 | hCoV-19/USA/WI-WSLH-200587/2020\|EPI_ISL_631389\|202 | ···················· | ···················· | ···················· | ···················· | NNNNNNNNNNNNNNNNNNNN | NNNNNNNNNNNNNNNNNNNN | 1 |
| Q | 7767 | hCoV-19/USA/WI-WSLH-200588/2020\|EPI_ISL_631390\|202 | ···················· | ···················· | ···················· | ···················· | NNNNNNNNNNNNNNNNNNNN | NNNNNNNNNNNNNNNNNNNN | 1 |
| Q | 7768 | hCoV-19/USA/WI-WSLH-200589/2020\|EPI_ISL_631391\|202 | ···················· | ···················· | ···················· | ···················· | NNNNNNNNNNNNNNNNNNNN | NNNNNNNNNNNNNNNNNNNN | 1 |
| Q | 7769 | hCoV-19/USA/WI-WSLH-200590/2020\|EPI_ISL_631392\|202 | ···················· | ···················· | ···················· | ···················· | NNNNNNNNNNNNNNNNNNNN | NNNNNNNNNNNNNNNNNNNN | 1 |
| Q | 7770 | hCoV-19/USA/WI-WSLH-200591/2020\|EPI_ISL_631393\|202 | ···················· | ···················· | ···················· | ···················· | NNNNNNNNNNNNNNNNNNNN | NNNNNNNNNNNNNNNNNNNN | 1 |
| Q | 7771 | hCoV-19/USA/WI-WSLH-200592/2020\|EPI_ISL_631394\|202 | ···················· | ···················· | ···················· | ···················· | NNNNNNNNNNNNNNNNNNNN | NNNNNNNNNNNNNNNNNNNN | 1 |
| Q | 7772 | hCoV-19/USA/WI-WSLH-200593/2020\|EPI_ISL_631395\|202 | ···················· | ···················· | ···················· | ···················· | NNNNNNNNNNNNNNNNNNNN | NNNNNNNNNNNNNNNNNNNN | 1 |
| Q | 7773 | hCoV-19/USA/WI-WSLH-200594/2020\|EPI_ISL_631396\|202 | ···················· | ···················· | ···················· | ···················· | NNNNNNNNNNNNNNNNNNNN | NNNNNNNNNNNNNNNNNNNN | 1 |
| Q | 7774 | hCoV-19/USA/WI-WSLH-200600/2020\|EPI_ISL_631397\|202 | ···················· | ···················· | ···················· | ···················· | NNNNNNNNNNNNNNNNNNNN | NNNNNNNNNNNNNNNNNNNN | 1 |
| Q | 7775 | hCoV-19/USA/WI-WSLH-200603/2020\|EPI_ISL_631398\|202 | ···················· | ···················· | ···················· | ···················· | NNNNNNNNNNNNNNNNNNNN | NNNNNNNNNNNNNNNNNNNN | 1 |
| Q | 7776 | hCoV-19/USA/WI-WSLH-200605/2020\|EPI_ISL_631399\|202 | ···················· | ···················· | ···················· | ···················· | NNNNNNNNNNNNNNNNNNNN | NNNNNNNNNNNNNNNNNNNN | 1 |
| Q | 7947 | hCoV-19/Wales/PHWC-4840E2/2020\|EPI_ISL_627965\|2020 | ···················· | ···················· | ···················· | ···················· | ··NNNNNNNNNNNNNNNNNN | ···················· | 1 |
| Q | 8222 | hCoV-19/Wales/PHWC-4840F1/2020\|EPI_ISL_627966\|2020 | ···················· | ···················· | ···················· | ···················· | ·NNNNNNNNNNNNNNNNN·· | ···················· | 1 |
| Q | 8224 | hCoV-19/Wales/PHWC-48413A/2020\|EPI_ISL_627969\|2020 | ···················· | ···················· | ···················· | ···················· | NNNNNNNNNNNNNNNNNNNN | NNNNNNNNNNNNNNNNNNNN | 1 |
| Q | 8241 | hCoV-19/Wales/PHWC-4842EC/2020\|EPI_ISL_627986\|2020 | ···················· | ···················· | ···················· | ···················· | NNNN················ | ···················· | 1 |
| Q | 8502 | hCoV-19/Wales/PHWC-483E43/2020\|EPI_ISL_627936\|2020 | ···················· | ···················· | ···················· | ···················· | NNNNNNNNNNNNNNNNNNNN | ···················· | 1 |
| Q | 8504 | hCoV-19/Wales/PHWC-483E61/2020\|EPI_ISL_627938\|2020 | ···················· | ···················· | ···················· | ···················· | NNNNNNNNNNNNNNNNNNNN | NNNNNNNNNNNNNNNNNNNN | 1 |
| Q | 8510 | hCoV-19/Wales/PHWC-483EE9/2020\|EPI_ISL_627944\|2020 | ···················· | ···················· | ···················· | ···················· | ···················· | NNNNNNNNNNNNNNNNNNNN | 1 |
| R | 257 | hCoV-19/Australia/VIC16563/2020\|EPI_ISL_641208\|202 | ···················· | ···················· | ···················· | ···················· | NNNNNNNNNNNNNNNNNN·· | NNNNNNNNNNNNNNNNNNNN | 1 |
| R | 261 | hCoV-19/Australia/VIC16302/2020\|EPI_ISL_641214\|202 | ···················· | ···················· | ···················· | ···················· | NNNNNNNNNNNNNNNNNNNN | ···················· | 1 |
| R | 263 | hCoV-19/Australia/VIC16368/2020\|EPI_ISL_641217\|202 | ···················· | ···················· | ···················· | ···················· | NNNNNNNNNNNNNNNNNNNN | NNNNNNNNNNNNNNNNNNNN | 1 |
| R | 273 | hCoV-19/Australia/VIC16615/2020\|EPI_ISL_641230\|202 | ···················· | ···················· | ···················· | ···················· | NNNNNNNNNNNNNNNNNNNN | ···················· | 1 |
| R | 282 | hCoV-19/Australia/VIC16334/2020\|EPI_ISL_641243\|202 | ···················· | ···················· | ···················· | ···················· | NNNNNNNNNNNNNNNNNNNN | ···················· | 1 |
| R | 283 | hCoV-19/Australia/VIC16325/2020\|EPI_ISL_641244\|202 | ···················· | ···················· | ···················· | ···················· | NNNNNNNNNNNNNNNNNNNN | NNNNNNNNNNNNNNNNNNNN | 1 |
| R | 289 | hCoV-19/Australia/VIC16693/2020\|EPI_ISL_641253\|202 | ···················· | ···················· | ···················· | ···················· | ·······NNNNNNNNNNNNN | ···················· | 1 |
| R | 292 | hCoV-19/Australia/VIC16681/2020\|EPI_ISL_641256\|202 | ···················· | ···················· | ···················· | ···················· | NNNNNNNNNNNNNNNNNNNN | NNNNNNNNNNNNNNNNNNNN | 1 |
| R | 299 | hCoV-19/Australia/VIC16293/2020\|EPI_ISL_641266\|202 | ···················· | ···················· | ···················· | ···················· | NNNNNNNNNNNNNNNNNNNN | ···················· | 1 |
| R | 307 | hCoV-19/Australia/VIC16545/2020\|EPI_ISL_641276\|202 | ···················· | ···················· | ···················· | ···················· | NNNNNNNNNNNNNNN····· | NNNN················ | 1 |
| R | 308 | hCoV-19/Australia/VIC16687/2020\|EPI_ISL_641278\|202 | ···················· | ···················· | ···················· | ···················· | ·············NNNN··· | ···················· | 1 |
| R | 513 | hCoV-19/Australia/VIC12336/2020\|EPI_ISL_641290\|202 | ···················· | ···················· | ···················· | ···················· | ·············NNNNNNN | ···················· | 1 |
| R | 775 | hCoV-19/Australia/VIC11779/2020\|EPI_ISL_641112\|202 | ···················· | ···················· | ···················· | ···················· | NNNNNNNNNNNNNNNNNNNN | NNNNNNNNNNNNNNNNNNNN | 1 |
| R | 803 | hCoV-19/Australia/VIC16673/2020\|EPI_ISL_641148\|202 | ···················· | ···················· | ···················· | ···················· | NNNNNNNNNNNNNNNNNNNN | NNNNNNNNNNNNNNNNNNNN | 1 |
| R | 1030 | hCoV-19/Australia/VIC16433/2020\|EPI_ISL_641172\|202 | ···················· | ···················· | ···················· | ···················· | NNNNNNNNNNNNNNNNN··· | NNNN····W··········· | 1 |
| R | 1039 | hCoV-19/Australia/VIC12635/2020\|EPI_ISL_641183\|202 | ···················· | ···················· | ···················· | ···················· | NNNNNNNNNNNNNNNNN··· | ···················· | 1 |
| R | 1041 | hCoV-19/Australia/VIC16159/2020\|EPI_ISL_641186\|202 | ···················· | ···················· | ···················· | ···················· | ···················· | NNNNNNNNN··········· | 1 |
| R | 1043 | hCoV-19/Australia/VIC16621/2020\|EPI_ISL_641189\|202 | ···················· | ···················· | ···················· | ···················· | NNNNNNNNNNNNNNNNNNNN | ···················· | 1 |
| R | 1050 | hCoV-19/Australia/VIC16471/2020\|EPI_ISL_641197\|202 | ···················· | ···················· | ···················· | ···················· | NNNNNNNNNNNNNNNNNNNN | NNN················· | 1 |
| R | 1290 | hCoV-19/Australia/VIC16281/2020\|EPI_ISL_641012\|202 | ···················· | ···················· | ···················· | ···················· | NNNNNNNNNNNN········ | ···················· | 1 |
| R | 1291 | hCoV-19/Australia/VIC16597/2020\|EPI_ISL_641013\|202 | ···················· | ···················· | ···················· | ···················· | NNNNNNNNNNNNNNNNNNNN | ···················· | 1 |
| R | 1296 | hCoV-19/Australia/VIC16679/2020\|EPI_ISL_641021\|202 | ···················· | ···················· | ···················· | ···················· | NNNNNNNNNNNNNNNNNNNN | NNNNN··············· | 1 |
| R | 1300 | hCoV-19/Australia/VIC16273/2020\|EPI_ISL_641026\|202 | ···················· | ···················· | ···················· | ···················· | NNNNNNNNNNNNNNNNNNNN | NNNNNNNNNNNNNNNNNNNN | 1 |
| R | 1302 | hCoV-19/Australia/VIC16674/2020\|EPI_ISL_641028\|202 | ···················· | ···················· | ···················· | ···················· | NNNNNNNNNNNNNNNNNNNN | ······NNNNNNNNN····· | 1 |
| R | 1525 | hCoV-19/USA/CA-CZB-10312/2020\|EPI_ISL_648503\|2020- | ···················· | ·A·················· | ···················· | ···················· | ···················· | NNNNNNNNNNNNNNN····· | 1 |
| R | 1539 | hCoV-19/Australia/VIC16454/2020\|EPI_ISL_641053\|202 | ···················· | ···················· | ···················· | ···················· | NNNNNNNNNNNNNNNNNNNN | NNNNNNNNNNNNNNNNNNNN | 1 |
| R | 1540 | hCoV-19/Australia/VIC16247/2020\|EPI_ISL_641054\|202 | ···················· | ···················· | ···················· | ···················· | ·NNNNNNNNNNNNNNN···· | ···················· | 1 |
| R | 1541 | hCoV-19/Australia/VIC16678/2020\|EPI_ISL_641056\|202 | ···················· | ···················· | ···················· | ···················· | NNNNNNN············· | ···················· | 1 |
| R | 1726 | hCoV-19/USA/CA-CZB-12333/2020\|EPI_ISL_648410\|2020- | ···················· | ·A·················· | ···················· | ···················· | NNNN················ | ···················· | 1 |
| R | 1727 | hCoV-19/USA/CA-CZB-12334/2020\|EPI_ISL_648411\|2020- | ···················· | ·A·················· | ···················· | ···················· | NNNNNNNNNNNNNNNN···· | ···················· | 1 |
| R | 1731 | hCoV-19/USA/CA-CZB-12344/2020\|EPI_ISL_648418\|2020- | ···················· | ·A·················· | ···················· | ···················· | NNNNNNNNN··········· | ···················· | 1 |
| R | 1761 | hCoV-19/USA/CA-CZB-12796/2020\|EPI_ISL_648451\|2020- | ···················· | ·A·················· | ···················· | ···················· | NNNNNNNNNNNNNNNNNNNN | ···NNNNNNNNNNNNNNNNN | 1 |
| R | 1763 | hCoV-19/USA/CA-CZB-12800/2020\|EPI_ISL_648453\|2020- | ···················· | ·A·················· | ···················· | ···················· | NNNNNNNNNNNNNNNNNNNN | ···················· | 1 |
| R | 1768 | hCoV-19/USA/CA-CZB-12809/2020\|EPI_ISL_648460\|2020- | ···················· | NNNNNNNNNNNNNNNNNNNN | ···················· | ···················· | ···················· | ···················· | 1 |
| R | 1769 | hCoV-19/USA/CA-CZB-12810/2020\|EPI_ISL_648461\|2020- | ···················· | ···················· | ···················· | ···················· | NNNNNNNNNNNNNNNNNNNN | NNNNNNNNNNNNNNNNNNNN | 1 |
| R | 2043 | hCoV-19/USA/CA-CZB-12847/2020\|EPI_ISL_648492\|2020- | ···················· | ·A·················· | ···················· | ···················· | NNNNNNNNNNNNNNNNNNNN | NNNNNNNNNNNNNNNNNNNN | 1 |
| R | 2047 | hCoV-19/USA/CA-CZB-7129/2020\|EPI_ISL_648496\|2020-0 | ···················· | ·A·················· | ···················· | ···················· | NNNNNNNNNNNNNNNNNNNN | NNNNNNNNNNNNNNNNNNNN | 1 |
| R | 2050 | hCoV-19/Australia/VIC16607/2020\|EPI_ISL_640359\|202 | ···················· | ···················· | ···················· | ···················· | NNNNNNNNNNNNNNNNNNNN | NNNNNNNNNNNNNNNNNNNN | 1 |
| R | 2053 | hCoV-19/Australia/VIC16381/2020\|EPI_ISL_640362\|202 | ···················· | ···················· | ···················· | ···················· | NNNNNNNNNNNNNNNNNNNN | NNNNNNNNNNNNNNNNNNNN | 1 |
| R | 2060 | hCoV-19/Australia/VIC16234/2020\|EPI_ISL_640385\|202 | ···················· | ···················· | ···················· | ···················· | NNNNNNNNNNNNNNNNNNNN | NNNNNNNNNNNNNNNNNNNN | 1 |
| R | 2070 | hCoV-19/Australia/VIC16648/2020\|EPI_ISL_640401\|202 | ···················· | ···················· | ···················· | ···················· | NNNNNNNNNNNNNNNNNNNN | NNNNNNNNNNNNNNNNNNNN | 1 |
| R | 2075 | hCoV-19/Australia/VIC16601/2020\|EPI_ISL_640406\|202 | ···················· | ···················· | ···················· | ···················· | NNNNNNNNNNNNNNNNNNN· | ···················· | 1 |
| R | 2076 | hCoV-19/Australia/VIC16691/2020\|EPI_ISL_640407\|202 | ···················· | ···················· | ···················· | ···················· | NNNNNNNNNNNNNNNNNNNN | NNNNNNNNNNNNNNNNNNNN | 1 |
| R | 2088 | hCoV-19/Australia/VIC16066/2020\|EPI_ISL_640422\|202 | ···················· | ···················· | ···················· | ···················· | NNNNNNNNNNNNNNNNNNNN | NNNNNNNNNNNNNNNN···· | 1 |
| R | 2106 | hCoV-19/Australia/VIC16387/2020\|EPI_ISL_640448\|202 | ···················· | ···················· | ···················· | ···················· | NNNNNNNNNNNNNNNNNNNN | ···················· | 1 |
| R | 2114 | hCoV-19/Australia/VIC16178/2020\|EPI_ISL_640457\|202 | ···················· | ···················· | ···················· | ···················· | NNNNNNNNNNNNNNNNNNNN | NNNNNNNNNNNNNNNNNNNN | 1 |
| R | 2306 | hCoV-19/Australia/VIC15998/2020\|EPI_ISL_640364\|202 | ···················· | ···················· | ···················· | ···················· | NNNNNNNNNNNNNNNNNNNN | ···················· | 1 |
| R | 2574 | hCoV-19/Australia/VIC16235/2020\|EPI_ISL_640304\|202 | ···················· | ···················· | ···················· | ···················· | NNNNNNNNNNNNNNNNNNNN | NNNNNNNNNNNNNNNNNNNN | 1 |
| R | 2578 | hCoV-19/Australia/VIC12537/2020\|EPI_ISL_640312\|202 | ···················· | ···················· | ···················· | ···················· | NNNNNNNNNNNNNNNNNNNN | ···················· | 1 |
| R | 2583 | hCoV-19/Australia/VIC16457/2020\|EPI_ISL_640317\|202 | ···················· | ···················· | ···················· | ···················· | NNNNNNNNNNNNNNNNNNNN | ···················· | 1 |
| R | 2585 | hCoV-19/Australia/VIC16312/2020\|EPI_ISL_640320\|202 | ···················· | ···················· | ···················· | ···················· | NNNNNNNNNNNNNNNNNNNN | NNNNNNNNNNNNNNNNNNNN | 1 |
| R | 2600 | hCoV-19/Australia/VIC16546/2020\|EPI_ISL_640340\|202 | ···················· | ···················· | ···················· | ···················· | NNNNNNNNNNNNNNNNN··· | ···················· | 1 |
| R | 2607 | hCoV-19/Australia/VIC16431/2020\|EPI_ISL_640352\|202 | ···················· | ···················· | ···················· | ···················· | NNNNNNNNNNNNNNNNNNNN | ···················· | 1 |
| R | 2608 | hCoV-19/Australia/VIC12105/2020\|EPI_ISL_640353\|202 | ···················· | ···················· | ···················· | ···················· | NNNNNNNNNNNNNNNNNNNN | ···················· | 1 |

| | | | | | | | | |
|---|---|---|---|---|---|---|---|---|
| R | 2613 hCoV-19/Belgium/ULG-10748/2020\|EPI_ISL_641590\|2020 | ···················· | ···················· | ···················· | ···················· | NNNNNNNNNNNNNNNNNNNN | NNNNNNNNNNNNNNNNNNNN | 1 |
| R | 2617 hCoV-19/Belgium/ULG-10752/2020\|EPI_ISL_641594\|2020 | ···················· | ···················· | ···················· | ···················· | NNNNNNNNNNNNNNNNNNNN | NNNNNNNNNNNNNNNNNNNN | 1 |
| R | 2618 hCoV-19/Belgium/ULG-10753/2020\|EPI_ISL_641595\|2020 | ···················· | ···················· | ···················· | ···················· | NNNNNNNNNNNNNNNNNNNN | NNNNNNNNNNNNNNNNNNNN | 1 |
| R | 2619 hCoV-19/Belgium/ULG-10754/2020\|EPI_ISL_641596\|2020 | ···················· | ···················· | ···················· | ···················· | NNNNNNNNNNNNNNNNNNNN | NNNNNNNNNNNNNNNNNNNN | 1 |
| R | 2622 hCoV-19/Belgium/ULG-10759/2020\|EPI_ISL_641601\|2020 | ···················· | ···················· | ···················· | ···················· | NNNNNNNNNNNNNNNNNNNN | NNNNNNNNNNNNNNNNNNNN | 1 |
| R | 2623 hCoV-19/Belgium/ULG-10760/2020\|EPI_ISL_641602\|2020 | ···················· | ···················· | ···················· | ···················· | NNNNNNNNNNNNNNNNNNNN | NNNNNNNNNNNNNNNNNNNN | 1 |
| R | 2625 hCoV-19/Belgium/ULG-10763/2020\|EPI_ISL_641605\|2020 | ···················· | ···················· | ···················· | ···················· | NNNNNNNNNNNNNNNNNNNN | NNNNNNNNNNNNNNNNNNNN | 1 |
| R | 2626 hCoV-19/Belgium/ULG-10764/2020\|EPI_ISL_641606\|2020 | ···················· | ···················· | ···················· | ···················· | NNNNNNNNNNNNNNNNNNNN | NNNNNNNNNNNNNNNNNNNN | 1 |
| R | 2838 hCoV-19/Germany/BW-MVZ-LS-55/2020\|EPI_ISL_640272\|2 | ···················· | ···················· | ···················· | ···················· | NNNNNNNNNNNNNNNNNNNN | NNNNNNNNNNNNNNNNNNNN | 1 |
| R | 2839 hCoV-19/Belgium/ULG-10724/2020\|EPI_ISL_641566\|2020 | ···················· | ···················· | ···················· | ···················· | NNNNNNNNNNNNNNNNNNNN | NNNNNNNNNNNNNNNNNNNN | 1 |
| R | 2840 hCoV-19/Belgium/ULG-10725/2020\|EPI_ISL_641567\|2020 | ···················· | ···················· | ···················· | ···················· | NNNNNNNNNNNNNNNNNNNN | NNNNNNNNNNNNNNNNNNNN | 1 |
| R | 2842 hCoV-19/Belgium/ULG-10727/2020\|EPI_ISL_641569\|2020 | ···················· | ···················· | ···················· | ···················· | NNNNNNNNNNNNNNNNNNNN | NNNNNNNNNNNNNNNNNNNN | 1 |
| R | 2843 hCoV-19/Belgium/ULG-10728/2020\|EPI_ISL_641570\|2020 | ···················· | ···················· | ···················· | ···················· | NNNNNNNNNNNNNNNNNNNN | NNNNNNNNNNNNNNNNNNNN | 1 |
| R | 2844 hCoV-19/Belgium/ULG-10729/2020\|EPI_ISL_641571\|2020 | ···················· | ···················· | ···················· | ···················· | NNNN················ | ···················· | 1 |
| R | 2845 hCoV-19/Belgium/ULG-10730/2020\|EPI_ISL_641572\|2020 | ···················· | ···················· | ···················· | ···················· | NNNNNNNNNNNNNNNNNNNN | NNNNNNNNNNNNNNNNNNNN | 1 |
| R | 2846 hCoV-19/Belgium/ULG-10731/2020\|EPI_ISL_641573\|2020 | ···················· | ···················· | ···················· | ···················· | NNNNNNNNNNNNNNNNNNNN | NNNNNNNNNNNNNNNNNNNN | 1 |
| R | 2847 hCoV-19/Belgium/ULG-10732/2020\|EPI_ISL_641574\|2020 | ···················· | ···················· | ···················· | ···················· | NNNNNNNNNNNNNNNNNNNN | NNNNNNNNNNNNNNNNNNNN | 1 |
| R | 2848 hCoV-19/Belgium/ULG-10733/2020\|EPI_ISL_641575\|2020 | ···················· | ···················· | ···················· | ···················· | NNNNNNNNNNNNNNNNNNNN | NNNNNNNNNNNNNNNNNNNN | 1 |
| R | 2849 hCoV-19/Belgium/ULG-10734/2020\|EPI_ISL_641576\|2020 | ···················· | ···················· | ···················· | ···················· | NNNNNNNNNNNNNNNNNNNN | NNNNNNNNNNNNNNNNNNNN | 1 |
| R | 2852 hCoV-19/Belgium/ULG-10737/2020\|EPI_ISL_641579\|2020 | ···················· | ···················· | ···················· | ···················· | NNNNNNNNNNNNNNNNNNNN | NNNNNNNNNNNNNNNNNNNN | 1 |
| R | 2853 hCoV-19/Belgium/ULG-10738/2020\|EPI_ISL_641580\|2020 | ···················· | ···················· | ···················· | ···················· | NNNNNNNNNNNNNNNNNNNN | NNNNNNNNNNNNNNNNNNNN | 1 |
| R | 2855 hCoV-19/Belgium/ULG-10741/2020\|EPI_ISL_641583\|2020 | ···················· | ···················· | ···················· | ···················· | NNNNNNNNNNNNNNNNNNNN | NNNNNNNNNNNNNNNNNNNN | 1 |
| R | 2857 hCoV-19/Belgium/ULG-10743/2020\|EPI_ISL_641585\|2020 | ···················· | ···················· | ···················· | ···················· | NNNNNNNNNNNNNNNNNNNN | NNNNNNNNNNNNNNNNNNNN | 1 |
| R | 2860 hCoV-19/Belgium/ULG-10757/2020\|EPI_ISL_641599\|2020 | ···················· | ·A·················· | ···················· | ···················· | NNNNNNNNNNNNNNNNNNNN | NNNNNNNNNNNNNNNNNNNN | 1 |
| R | 3073 hCoV-19/USA/MI-UM-10036558900/2020\|EPI_ISL_640205\| | ···················· | ·····A·············· | ···················· | ···················· | NNNNNNNNNNNNNNNNNNNN | ···················· | 1 |
| R | 3074 hCoV-19/USA/MI-UM-10036558247/2020\|EPI_ISL_640206\| | ···················· | ···················· | ···················· | ···················· | NNNNNNNNNNNNNNNNNNNN | NNNNNNNNNNNNNNNNNNNN | 1 |
| R | 3075 hCoV-19/USA/MI-UM-10036558969/2020\|EPI_ISL_640207\| | ···················· | ·····A·············· | ···················· | ···················· | NNNNNNNNNNNNNNNNNNNN | ···················· | 1 |
| R | 3076 hCoV-19/USA/MI-UM-10036571832/2020\|EPI_ISL_640208\| | ···················· | ···················· | ···················· | ···················· | NNNNNNNNNNNNNNNNNNNN | NNNNNNNNNNNNNNNNNNNN | 1 |
| R | 3077 hCoV-19/USA/MI-UM-10036556633/2020\|EPI_ISL_640209\| | ···················· | ···················· | ···················· | ···················· | NNNNNNNNNNNNNNNNNNNN | NNNNNNNNNNNNNNNNNNNN | 1 |
| R | 3078 hCoV-19/USA/MI-UM-10036567026/2020\|EPI_ISL_640211\| | ···················· | ·A·················· | ···················· | ···················· | NNNNNNNNNNNNNNNNNNNN | ···················· | 1 |
| R | 3080 hCoV-19/USA/MI-UM-10036568766/2020\|EPI_ISL_640214\| | ···················· | ·············A······ | ···················· | ···················· | NNNNNNNNNNNNNNNNNNNN | ···················· | 1 |
| R | 3082 hCoV-19/USA/MI-UM-10036605690/2020\|EPI_ISL_640216\| | ···················· | ···················· | ···················· | ···················· | NNNNNNNNNNNNNNNNNNNN | NNNNNNNNNNNNNNNNNNNN | 1 |
| R | 3083 hCoV-19/USA/MI-UM-10036591651/2020\|EPI_ISL_640218\| | ···················· | ···················· | ···················· | ···················· | NNNNNNNNNNNNNNNNNNNN | NNNNNNNNNNNNNNNNNNNN | 1 |
| R | 3136 hCoV-19/Belgium/ULG-10716/2020\|EPI_ISL_641558\|2020 | ···················· | ···················· | ···················· | ···················· | NNNNNNNNNNNNNNNNNNNN | NNNNNNNNNNNNNNNNNNNN | 1 |
| R | 3137 hCoV-19/Belgium/ULG-10717/2020\|EPI_ISL_641559\|2020 | ···················· | ···················· | ···················· | ···················· | NNNNNNNNNNNNNNNNNNNN | NNNNNNNNNNNNNNNNNNNN | 1 |
| R | 3138 hCoV-19/Belgium/ULG-10718/2020\|EPI_ISL_641560\|2020 | ···················· | ···················· | ···················· | ···················· | NNNNNNNNNNNNNNNNNNNN | NNNNNNNNNNNNNNNNNNNN | 1 |
| R | 3139 hCoV-19/Belgium/ULG-10719/2020\|EPI_ISL_641561\|2020 | ···················· | ···················· | ···················· | ···················· | NNNNNNNNNNNNNNNNNNNN | NNNNNNNNNNNNNNNNNNNN | 1 |
| R | 3140 hCoV-19/Belgium/ULG-10720/2020\|EPI_ISL_641562\|2020 | ···················· | ···················· | ···················· | ···················· | NNNNNNNNNNNNNNNNNNNN | NNNNNNNNNNNNNNNNNNNN | 1 |
| R | 3141 hCoV-19/Belgium/ULG-10721/2020\|EPI_ISL_641563\|2020 | ···················· | ···················· | ···················· | ···················· | NNNNNNNNNNNNNNNNNNNN | NNNNNNNNNNNNNNNNNNNN | 1 |
| R | 3143 hCoV-19/Belgium/ULG-10723/2020\|EPI_ISL_641565\|2020 | ···················· | ···················· | ···················· | ···················· | NNNNNNNNNNNNNNNNNNNN | NNNNNNNNNNNNNNNNNNNN | 1 |
| R | 3208 hCoV-19/England/MILK-B01E73/2020\|EPI_ISL_642896\|20 | ···················· | ···················· | ···················· | NNNNNNNNNNNNNNNNNNNN | ···················· | ···················· | 1 |
| R | 3352 hCoV-19/USA/MI-UM-10036540781/2020\|EPI_ISL_640146\| | ···················· | ···················· | ···················· | ···················· | NNNNNNNNNNNNNNNNNNNN | ···················· | 1 |
| R | 3353 hCoV-19/USA/MI-UM-10036530676/2020\|EPI_ISL_640147\| | ···················· | ···················· | ···················· | ···················· | NNNNNNNNNNNNNNNNNNNN | ···················· | 1 |
| R | 3354 hCoV-19/USA/MI-UM-10036535365/2020\|EPI_ISL_640148\| | ···················· | ·····A·············· | ···················· | ···················· | ·······NNNNNNNNNNNNN | ···················· | 1 |
| R | 3359 hCoV-19/USA/MI-UM-10036535709/2020\|EPI_ISL_640153\| | ···················· | ·A·················· | ···················· | ···················· | NNNNNNNNNNNNNNNNNNNN | ···················· | 1 |
| R | 3368 hCoV-19/USA/MI-UM-10036578565/2020\|EPI_ISL_640163\| | ···················· | ···················· | ···················· | ···················· | NNNNNNNNNNNNNNNNNNNN | ···················· | 1 |
| R | 3369 hCoV-19/USA/MI-UM-10036557572/2020\|EPI_ISL_640164\| | ···················· | ·············A······ | ···················· | ···················· | NNNNNNNNNNNNNNNNNNNN | ···················· | 1 |
| R | 3385 hCoV-19/USA/MI-UM-10036556732/2020\|EPI_ISL_640182\| | ···················· | ···················· | ···················· | ···················· | NNNNNNNNNNNNNNNNNNNN | NNNNNNNNNNNNNNNNNNNN | 1 |
| R | 3386 hCoV-19/USA/MI-UM-10036578191/2020\|EPI_ISL_640183\| | ···················· | ·A·················· | ···················· | ···················· | NNNNNNNNNNNNNNNNNNNN | ···················· | 1 |
| R | 3387 hCoV-19/USA/MI-UM-10036561289/2020\|EPI_ISL_640184\| | ···················· | ···················· | ···················· | ···················· | NNNNNNNNNNNNNNNNNNNN | NNNNNNNNNNNNNNNNNNNN | 1 |
| R | 3388 hCoV-19/USA/MI-UM-10036548240/2020\|EPI_ISL_640185\| | ···················· | ·A·················· | ···················· | ···················· | NNNNNNNNNNNNNNNNNNNN | ···················· | 1 |
| R | 3389 hCoV-19/USA/MI-UM-10036577344/2020\|EPI_ISL_640186\| | ···················· | ···················· | ···················· | ···················· | NNNNNNNNNNNNNNNNNNNN | NNNNNNNNNNNNNNNNNNNN | 1 |
| R | 3392 hCoV-19/USA/MI-UM-10036541021/2020\|EPI_ISL_640192\| | ···················· | ···················· | ···················· | ···················· | NNNNNNNNNNNNNNNNNNNN | NNNNNNNNNNNNNNNNNNNN | 1 |
| R | 3393 hCoV-19/USA/MI-UM-10036556206/2020\|EPI_ISL_640193\| | ···················· | ···················· | ···················· | ···················· | NNNNNNNNNNNNNNNNNNNN | ···················· | 1 |
| R | 3394 hCoV-19/USA/MI-UM-10036558807/2020\|EPI_ISL_640195\| | ···················· | ···················· | ···················· | ···················· | NNNNNNNNNNNNNNNNNNNN | NNNNNNNNNNNNNNNNNNNN | 1 |
| R | 3395 hCoV-19/USA/MI-UM-10036551324/2020\|EPI_ISL_640196\| | ···················· | ···················· | ···················· | ···················· | NNNNNNNNNNNNNNNNNNNN | NNNNNNNNNNNNNNNNNNNN | 1 |
| R | 3592 hCoV-19/mink/Denmark/mDK-26/2020\|EPI_ISL_641419\|20 | ···················· | ····A··············· | ···················· | ···················· | NNNNNNNNNNNNNNNNNNNN | NNNNNNNNNNNNNNNNNNNN | 1 |
| R | 3593 hCoV-19/mink/Denmark/mDK-27/2020\|EPI_ISL_641420\|20 | ···················· | ····A··C············ | ···················· | ···················· | NNNNNNNNNNNNNNNNNNNN | NNNNNNNNNNNNNNNNNNNN | 1 |
| R | 3594 hCoV-19/mink/Denmark/mDK-28/2020\|EPI_ISL_641421\|20 | ···················· | ···················· | ···················· | ···················· | NNNNNNNNNNNNNNNNNNNN | NNNNNNNNNNNNNNNNNNNN | 1 |
| R | 3596 hCoV-19/mink/Denmark/mDK-30/2020\|EPI_ISL_641423\|20 | ···················· | ···················· | ···················· | ···················· | NNNNNNNNNNNNNNNNNNNN | NNNNNNNNNNNNNNNNNNNN | 1 |
| R | 3874 hCoV-19/South Africa/NHLS-UCT-GS-0022/2020\|EPI_ISL | ···················· | ···················· | ···················· | ···················· | ··NNNNNNNNNNNNNNNNNN | ···················· | 1 |
| R | 3902 hCoV-19/South Africa/NHLS-UCT-GS-3555/2020\|EPI_ISL | ···················· | ·A·················· | ···················· | ···················· | NNNN················ | ···················· | 1 |
| R | 3903 hCoV-19/Australia/VIC16532/2020\|EPI_ISL_640851\|202 | ···················· | ···················· | ···················· | ···················· | NNNNNNNNNNNNNNNNNNNN | ···················· | 1 |
| R | 4189 hCoV-19/Australia/VIC16696/2020\|EPI_ISL_640824\|202 | ···················· | ···················· | ···················· | ···················· | NNNNNNNNNNNNNNNNNNNN | NNNNNNNNNNNNNNNNNNNN | 1 |
| R | 4192 hCoV-19/Australia/VIC16508/2020\|EPI_ISL_640829\|202 | ···················· | ···················· | ···················· | ···················· | NNNNNNNNNNNNNNNNNNNN | ···················· | 1 |
| R | 4194 hCoV-19/Australia/VIC16517/2020\|EPI_ISL_640831\|202 | ···················· | ···················· | ···················· | ···················· | NNNNNNNNNNNNNNNNNNNN | ···················· | 1 |
| R | 4219 hCoV-19/Australia/VIC16668/2020\|EPI_ISL_640876\|202 | ···················· | ···················· | ···················· | ···················· | NNNNNNNNNNNNNNNNNNNN | ············NNNNNNNN | 1 |
| R | 4226 hCoV-19/Australia/VIC16415/2020\|EPI_ISL_640883\|202 | ···················· | ···················· | ···················· | ···················· | NNNNNNNNNNNNNNNNNNNN | NNNNNNNNNNNNNNNNNNNN | 1 |
| R | 4239 hCoV-19/Australia/VIC16533/2020\|EPI_ISL_640896\|202 | ···················· | ···················· | ···················· | ···················· | NNNNNNNNNNNNNNNNNNNN | NN·················· | 1 |
| R | 4245 hCoV-19/England/QEUH-A8B434/2020\|EPI_ISL_643073\|20 | ···················· | ···················· | ···················· | NNNNNNNNNNNNNNNNNNNN | ···················· | ···················· | 1 |
| R | 4376 hCoV-19/Australia/VIC16122/2020\|EPI_ISL_640745\|202 | ···················· | ···················· | ···················· | ···················· | NNNNN··············· | ···················· | 1 |
| R | 4659 hCoV-19/Australia/VIC16447/2020\|EPI_ISL_640704\|202 | ···················· | ···················· | ···················· | ···················· | NNNNNNNNNNNNNNNNNNNN | ···················· | 1 |
| R | 4672 hCoV-19/Australia/VIC16711/2020\|EPI_ISL_640721\|202 | ···················· | ···················· | ···················· | ···················· | NNNNNNNNNNNNNNNNNNNN | NNNNNNNNNNNNNNNN···· | 1 |
| R | 4680 hCoV-19/Australia/VIC16274/2020\|EPI_ISL_640753\|202 | ···················· | ···················· | ···················· | ···················· | NNNNNNNNNNNNNNNNNNNN | NNNNNNNNNNNNNNNNNNNN | 1 |
| R | 4681 hCoV-19/Australia/VIC16219/2020\|EPI_ISL_640754\|202 | ···················· | ···················· | ···················· | ···················· | NNNNNNNNNNNN········ | ···················· | 1 |
| R | 4694 hCoV-19/Australia/VIC12885/2020\|EPI_ISL_640776\|202 | ···················· | ········W··········· | ···················· | ···················· | NNNNNNNNNNNNNNNNNNNN | NNNNNNNNNNNNNNNNNNNN | 1 |
| R | 4697 hCoV-19/Australia/VIC16672/2020\|EPI_ISL_640780\|202 | ···················· | ···················· | ···················· | ···················· | NNNNNNNNNNNNNNNNNNNN | NNNNNNNNNNNNNNNNNNNN | 1 |
| R | 4698 hCoV-19/Australia/VIC12560/2020\|EPI_ISL_640781\|202 | ···················· | ·····A·············· | ···················· | ···················· | NNNNNNNNNNNNNNNNNNNN | NNNNNNNNNNNNNNNNNNNN | 1 |
| R | 4813 hCoV-19/USA/MI-UM-10036798275/2020\|EPI_ISL_649057\| | ···················· | ·A·················· | ···················· | ···················· | NNNNNNNNNNNNNNNNNNNN | NNNNNNNNNNNNNNNNNNNN | 1 |
| R | 4814 hCoV-19/USA/MI-UM-10036728634/2020\|EPI_ISL_649058\| | ···················· | ·A·················· | ···················· | ···················· | NNNNNNNNNNNNNNNNNNNN | NNNNNNNNNNNNNNNNNNNN | 1 |

| | | | | | | | | |
|---|---|---|---|---|---|---|---|---|
| R | 4815 hCoV-19/USA/MI-UM-10036787455/2020\|EPI_ISL_649059\| | ···················· | ·A·················· | ···················· | ···················· | NNNNNNNNNNNNNNNNNNNN | NNNNNNNNNNNNNNNNNNNN | 1 |
| R | 4834 hCoV-19/Israel/CVL-n4326/2033\|EPI_ISL_649084\|2020- | ···················· | ···················· | ···················· | ···················· | ········NNNNNNNNNNNN | ···················· | 1 |
| R | 4882 hCoV-19/Australia/VIC16549/2020\|EPI_ISL_640628\|202 | ···················· | ···················· | ···················· | ···················· | NNNNNNNNNNNNNNNNNNNN | NNNNNNNNNNNNNNNNNNNN | 1 |
| R | 4883 hCoV-19/Australia/VIC13062/2020\|EPI_ISL_640629\|202 | ···················· | ···················· | ···················· | ···················· | NNNNNNNNNNNNNNNNNNNN | ···················· | 1 |
| R | 4976 hCoV-19/Ireland/D-NVRL-AIIDV0083v1/2020\|EPI_ISL_64 | ···················· | ···················· | ···················· | ···················· | NNNNNNNNNNNNNNNNNNNN | NNNNNNNNNNNNNNNNNNNN | 1 |
| R | 4981 hCoV-19/Ireland/D-NVRL-AIIDV0432v2/2020\|EPI_ISL_64 | ···················· | ···················· | ···················· | ···················· | NNNNNNNNNNNNNNNNNNNN | NNNNNNNNNNNNNNNNNNNN | 1 |
| R | 5127 hCoV-19/Australia/VIC16688/2020\|EPI_ISL_640637\|202 | ···················· | ···················· | ···················· | ···················· | NNNNNNNNNNNNNNNNNNNN | NNNNNNNNNNNNNNNNNNNN | 1 |
| R | 5130 hCoV-19/Australia/VIC16081/2020\|EPI_ISL_640643\|202 | ···················· | ···················· | ···················· | ···················· | NNNNNNNNNNNN········ | ···················· | 1 |
| R | 5135 hCoV-19/Australia/VIC12838/2020\|EPI_ISL_640651\|202 | ···················· | ···················· | ···················· | ···················· | NNNNNNNNNNNNNNNNNNNN | NNNNNNNNNNNNNNNNNNNN | 1 |
| R | 5142 hCoV-19/Australia/VIC16712/2020\|EPI_ISL_640660\|202 | ···················· | ···················· | ···················· | ···················· | NNNNNNNNNNNNNNNNNNNN | NNNNNNNNNNNNNNNNNNNN | 1 |
| R | 5144 hCoV-19/Australia/VIC16272/2020\|EPI_ISL_640662\|202 | ···················· | ···················· | ···················· | ···················· | NNNNNNNNNNNNNNNNNNNN | NNNNNNNNNNNNNNNNNNNN | 1 |
| R | 5147 hCoV-19/Australia/VIC12242/2020\|EPI_ISL_640666\|202 | ···················· | ···················· | ···················· | ···················· | NNNNNNNNNNNNNNNNNNNN | NNNNNNNNNNNNNNNNNNNN | 1 |
| R | 5150 hCoV-19/Australia/VIC16614/2020\|EPI_ISL_640670\|202 | ···················· | ···················· | ···················· | ···················· | NNNNNNNNNNNNNNNNNNNN | NNNNNNNNNNNNNNNNNNNN | 1 |
| R | 5158 hCoV-19/Australia/VIC16021/2020\|EPI_ISL_640689\|202 | ···················· | ···················· | ···················· | ···················· | NNNNNNNNNNNN········ | ···················· | 1 |
| R | 5159 hCoV-19/Australia/VIC16263/2020\|EPI_ISL_640690\|202 | ···················· | ···················· | ···················· | ···················· | NNNNNNNNNNNNNNNNNNNN | ···················· | 1 |
| R | 5162 hCoV-19/Australia/VIC16014/2020\|EPI_ISL_640693\|202 | ···················· | ···················· | ···················· | ···················· | NNNNNNNNNNNNNNNNNNNN | ···················· | 1 |
| R | 5164 hCoV-19/Australia/VIC15963/2020\|EPI_ISL_640695\|202 | ···················· | ···················· | ···················· | ···················· | NNNNNNNNNNNNNNNNNNNN | ·R·················· | 1 |
| R | 5377 hCoV-19/Australia/VIC12250/2020\|EPI_ISL_640500\|202 | ···················· | ···················· | ···················· | ···················· | NNNNNNNNNNNNNNNNNNNN | NNNNNNNNNNN········· | 1 |
| R | 5390 hCoV-19/Australia/VIC16703/2020\|EPI_ISL_640516\|202 | ···················· | ···················· | ···················· | ···················· | NNNNNNNNNNNNNNNNNNNN | NNNNNNNNNNNNNNNNNNNN | 1 |
| R | 5395 hCoV-19/Australia/VIC16369/2020\|EPI_ISL_640521\|202 | ···················· | ···················· | ···················· | ···················· | NNNNNNNNNNNNNNNNNNNN | NNNNNNNNNNNNNNNNNNNN | 1 |
| R | 5398 hCoV-19/Australia/VIC16367/2020\|EPI_ISL_640528\|202 | ···················· | ···················· | ···················· | ···················· | NNNNNNNNNNNNNNNNNNNN | ···················· | 1 |
| R | 5415 hCoV-19/Australia/VIC11985/2020\|EPI_ISL_640557\|202 | ···················· | ···················· | ···················· | ···················· | ···················· | ·NNNNNNNNNNN········ | 1 |
| R | 5417 hCoV-19/Australia/VIC16632/2020\|EPI_ISL_640559\|202 | ···················· | ···················· | ···················· | ···················· | NNNNNNNNNNNNNNNNNNNN | NNNNNNNNNNNNNNNNNNNN | 1 |
| R | 5420 hCoV-19/Australia/VIC16348/2020\|EPI_ISL_640565\|202 | ···················· | ···················· | ···················· | ···················· | NNNNNNNNNNNNNNNNNNNN | ···················· | 1 |
| R | 5425 hCoV-19/Australia/VIC16125/2020\|EPI_ISL_640571\|202 | ···················· | ···················· | ···················· | ···················· | ·············NNNNNNN | NNNN········NNNNNNNN | 1 |
| R | 5432 hCoV-19/Australia/VIC16695/2020\|EPI_ISL_640578\|202 | ···················· | ···················· | ···················· | ···················· | NNNNNNNNNNNNNNNNNNNN | NNNNNNNNNNNNNNNNNNNN | 1 |
| R | 5434 hCoV-19/Australia/VIC16054/2020\|EPI_ISL_640580\|202 | ···················· | ···················· | ···················· | ···················· | ······NNNNNN········ | ···················· | 1 |
| R | 5437 hCoV-19/Australia/VIC16303/2020\|EPI_ISL_640583\|202 | ···················· | ···················· | ···················· | ···················· | NNNNNNNNNNNNNNNNNNNN | NNNNNNNNNNNNNNNNNNNN | 1 |
| R | 5439 hCoV-19/Australia/VIC16269/2020\|EPI_ISL_640586\|202 | ···················· | ···················· | ···················· | ···················· | NNNNNNNNNNNNNNNNNNNN | ···················· | 1 |
| R | 5445 hCoV-19/Australia/VIC16053/2020\|EPI_ISL_640593\|202 | ···················· | ···················· | ···················· | ···················· | NNNNNNNNNNNNNNNNNNNN | ···················· | 1 |
| R | 5633 hCoV-19/Australia/VIC16371/2020\|EPI_ISL_640478\|202 | ···················· | ···················· | ···················· | ···················· | NNNNNNNNNNNNNNNNNNNN | ···················N | 1 |
| R | 5635 hCoV-19/Australia/VIC16261/2020\|EPI_ISL_640482\|202 | ···················· | ···················· | ···················· | ···················· | NNNNNNNNNNNNNNNNNNNN | NNNNNNNNNNNNNNNNNNNN | 1 |
| R | 5640 hCoV-19/Australia/VIC12067/2020\|EPI_ISL_640492\|202 | ···················· | ···················· | ···················· | ···················· | NNNNNNNNNNNNNNNNNNNN | ···················· | 1 |
| R | 5927 hCoV-19/Zimbabwe/ZW-1114/2020\|EPI_ISL_644786\|2020- | ···················· | ···················· | ···················· | ···················· | NNNNNNNNNNNNNNNNNNNN | NNNNNNNNNNNNNNNNNNNN | 1 |
| R | 5928 hCoV-19/Zimbabwe/ZW-25BY/2020\|EPI_ISL_644787\|2020- | ···················· | ···················· | ···················· | ···················· | NNNNNNNNNNNNNNNNNNNN | NNNNNNNNNNNNNNN····· | 1 |
| R | 5929 hCoV-19/Zimbabwe/ZW-28BY/2020\|EPI_ISL_644788\|2020- | ···················· | ···················· | ···················· | ···················· | NNNNNNNNNNNN········ | ···················· | 1 |
| R | 5930 hCoV-19/Zimbabwe/ZW-391/2020\|EPI_ISL_644789\|2020-0 | ···················· | ···················· | ···················· | ···················· | NNNNNNNNN··········· | ···················· | 1 |
| R | 5932 hCoV-19/Zimbabwe/ZW-6969/2020\|EPI_ISL_644791\|2020- | ···················· | ···················· | ···················· | ···················· | NNNNNNNNNNNNNNNNNNNN | NNNNNNNNNNNNNNNNNNNN | 1 |
| R | 5933 hCoV-19/Zimbabwe/ZW-6973/2020\|EPI_ISL_644792\|2020- | ···················· | ···················· | ···················· | ···················· | NNNNNNNNNNNNNNNNNNNN | ···················· | 1 |
| R | 5934 hCoV-19/Zimbabwe/ZW-6974/2020\|EPI_ISL_644793\|2020- | ···················· | ···················· | ···················· | ···················· | NNNNNNNNNNNNNNNNNNNN | NNNNNNNNNNNNNNNNNNNN | 1 |
| R | 5935 hCoV-19/Zimbabwe/ZW-6976/2020\|EPI_ISL_644794\|2020- | ···················· | ···················· | ···················· | ···················· | NNNNNNNNNNNNNNNNNNNN | NNNNNNNNNNNNNNNNNNNN | 1 |
| R | 5937 hCoV-19/Zimbabwe/ZW-6979/2020\|EPI_ISL_644796\|2020- | ···················· | ···················· | ···················· | ···················· | NNNNNNNNNNNNNNNNNNNN | NNNNNNNNNNNNNNNNNNNN | 1 |
| R | 5938 hCoV-19/Zimbabwe/ZW-6980/2020\|EPI_ISL_644797\|2020- | ···················· | ···················· | ···················· | ···················· | ···················· | NNNNN··············· | 1 |
| R | 5940 hCoV-19/Zimbabwe/ZW-6987/2020\|EPI_ISL_644799\|2020 | ···················· | ···················· | ···················· | ···················· | NNNNNNNNNNNNNNNNNNNN | NNNNNNNNNNNNNNNNNNNN | 1 |
| R | 5942 hCoV-19/Zimbabwe/ZW-6990/2020\|EPI_ISL_644801\|2020- | ···················· | ···················· | ···················· | ···················· | NNNNNNNNNNNNNNNNNNNN | ···················· | 1 |
| R | 5943 hCoV-19/Zimbabwe/ZW-6991/2020\|EPI_ISL_644802\|2020- | ···················· | ···················· | ···················· | ···················· | NNNNNNNNNNNNNNNNNNNN | NNNNNNNNNNNNNNNNNNNN | 1 |
| R | 5949 hCoV-19/Zimbabwe/ZW-6999/2020\|EPI_ISL_644808\|2020- | ···················· | ···················· | ···················· | ···················· | NNNN················ | ···················· | 1 |
| R | 5951 hCoV-19/Zimbabwe/ZW-7002/2020\|EPI_ISL_644810\|2020- | ···················· | ···················· | ···················· | ···················· | NNNNNNNNNNNNNNNNNNNN | ···················· | 1 |
| R | 5952 hCoV-19/Zimbabwe/ZW-7003/2020\|EPI_ISL_644811\|2020- | ···················· | ···················· | ···················· | ···················· | NNNN················ | ···················· | 1 |
| R | 5954 hCoV-19/Zimbabwe/ZW-9506/2020\|EPI_ISL_644813\|2020- | ···················· | ···················· | ···················· | ···················· | NNNNNNNNNNNNNNNNNNNN | ···················· | 1 |
| R | 5955 hCoV-19/Zimbabwe/ZW-9513/2020\|EPI_ISL_644814\|2020- | ···················· | ···················· | ···················· | ···················· | NNNN················ | ···················· | 1 |
| R | 5956 hCoV-19/Zimbabwe/ZW-9521/2020\|EPI_ISL_644815\|2020- | ···················· | ·A·················· | ···················· | ···················· | NNNNNNNNNNNNNNNNNNNN | NNNN················ | 1 |
| R | 5958 hCoV-19/Zimbabwe/ZW-9538/2020\|EPI_ISL_644817\|2020- | ···················· | ·A·················· | ···················· | ···················· | NNN················· | NNNNNNNNN··········· | 1 |
| R | 5960 hCoV-19/Zimbabwe/ZW-9949/2020\|EPI_ISL_644819\|2020- | ···················· | ···················· | ···················· | ···················· | NNNNNNNNNNNNNNNNNNNN | ···················· | 1 |
| R | 5962 hCoV-19/Zimbabwe/ZW-9955/2020\|EPI_ISL_644821\|2020- | ···················· | ···················· | ···················· | ···················· | NNNNNNNNNNNNNNNNNNNN | ···················· | 1 |
| R | 6090 hCoV-19/Equatorial Guinea/10798/2020\|EPI_ISL_64830 | ···················· | ···················· | ···················· | ···················· | NNNNNNNNNNNNNNNNNNNN | NNNNNNNNNNNNNNNNNNNN | 1 |
| R | 6091 hCoV-19/Equatorial Guinea/11514/2020\|EPI_ISL_64830 | ···················· | ···················· | ···················· | ···················· | NNNNNNNNNNNNNNNNNNNN | NNNN················ | 1 |
| R | 6092 hCoV-19/Equatorial Guinea/11678/2020\|EPI_ISL_64830 | ···················· | ···················· | ···················· | ···················· | NNNNNNNNNNNNNNNNNNNN | NNNNNNNNNNNNNNNNNNNN | 1 |
| R | 6093 hCoV-19/Equatorial Guinea/12170/2020\|EPI_ISL_64831 | ···················· | ···················· | ···················· | ···················· | NNNNNNNNNNNNNNNNNNNN | NNNNNNNNNNNNNNNNNNNN | 1 |
| R | 6095 hCoV-19/Equatorial Guinea/12864/2020\|EPI_ISL_64831 | ···················· | ···················· | ···················· | ···················· | NNNNNNNNNNNNNNNNNNNN | NNNNNNNNNNNNNNNNNNNN | 1 |
| R | 6097 hCoV-19/Equatorial Guinea/2682/2020\|EPI_ISL_648319 | ···················· | ···················· | ···················· | ···················· | NNNNNNNNNNNNNNNNNNNN | NNNNNNNNN··········· | 1 |
| R | 6098 hCoV-19/Equatorial Guinea/28098/2020\|EPI_ISL_64832 | ···················· | ···················· | ···················· | ···················· | NNNNNNNNNNNNNNNNNNNN | NNNNNNNNNNNNNNNNNNNN | 1 |
| R | 6099 hCoV-19/Equatorial Guinea/2892/2020\|EPI_ISL_648321 | ···················· | ···················· | ···················· | ···················· | NNNNNNNNNNNNNNNNNNNN | ···················· | 1 |
| R | 6100 hCoV-19/Equatorial Guinea/28974/2020\|EPI_ISL_64832 | ···················· | ···················· | ···················· | ···················· | NNNNNNNNNNNNNNNNNNNN | NNNNNNNNNNNNNNNNNNNN | 1 |
| R | 6101 hCoV-19/Equatorial Guinea/32447/2020\|EPI_ISL_64832 | ···················· | ···················· | ···················· | ···················· | NNNNNNNNNNNNNNNNNNNN | NNNNNNNNNNNNNNNNNNNN | 1 |
| R | 6102 hCoV-19/Equatorial Guinea/3280/2020\|EPI_ISL_648325 | ···················· | ···················· | ···················· | ···················· | NNNNNNNNNNNNNNNNNNNN | NNNNNNNNNNNNNNNNNNNN | 1 |
| R | 6104 hCoV-19/Equatorial Guinea/37030/2020\|EPI_ISL_64832 | ···················· | ···················· | ···················· | ···················· | NNNNNNNNNNNNNNNNNNNN | NNNN···NN··········· | 1 |
| R | 6105 hCoV-19/Equatorial Guinea/37310/2020\|EPI_ISL_64832 | ···················· | ···················· | ···················· | ···················· | NNNNNNNNNNNNNNNNNNNN | NNNNNNNNNNNNNNNNNNNN | 1 |
| R | 6106 hCoV-19/Equatorial Guinea/37545/2020\|EPI_ISL_64832 | ···················· | ···················· | ···················· | ···················· | NNNNNNNNNNNNNNNNNNNN | NNNNNNNNNNNNNNNNNNNN | 1 |
| R | 6107 hCoV-19/Equatorial Guinea/38087/2020\|EPI_ISL_64833 | ···················· | ···················· | ···················· | ···················· | NNNNNNNNNNNNNNNNNNNN | NNNNNNNNNNNNNNNNNNNN | 1 |
| R | 6108 hCoV-19/Equatorial Guinea/3910/2020\|EPI_ISL_648334 | ···················· | ···················· | ···················· | ···················· | NNNNNNNNNNNNNNNNNNNN | NNNNNNNNNNNNNNNNNNNN | 1 |
| R | 6109 hCoV-19/Equatorial Guinea/4634/2020\|EPI_ISL_648336 | ···················· | ···················· | ···················· | ···················· | NNNNNNNNNNNNNNNNNNNN | NNNNNNNNNNNNNNNNNNNN | 1 |
| R | 6110 hCoV-19/Equatorial Guinea/49849/2020\|EPI_ISL_64833 | ···················· | ···················· | ···················· | ···················· | NNNNNNNNNNNNNNNNNNNN | NNNNN··············· | 1 |
| R | 6111 hCoV-19/Equatorial Guinea/50236/2020\|EPI_ISL_64833 | ···················· | ···················· | ···················· | ···················· | NNNNNNNNNNNNNNNNNNNN | NNNNN··············· | 1 |
| R | 6114 hCoV-19/Equatorial Guinea/5508/2020\|EPI_ISL_648343 | ···················· | ···················· | ···················· | ···················· | NNNNNNNNNNNNNNNNNNNN | NNNNNNNNNNNNNNNNNNNN | 1 |
| R | 6116 hCoV-19/Equatorial Guinea/6818/2020\|EPI_ISL_648345 | ···················· | ···················· | ···················· | ···················· | NNNNNNNNNNNNNNNNNNNN | NNNNNNNNNNNNNNNNNNNN | 1 |
| R | 6117 hCoV-19/Equatorial Guinea/7692/2020\|EPI_ISL_648347 | ···················· | ···················· | ···················· | ···················· | NNNNNNNNNNNNNNNNNNNN | NNNNNNNNNNNNNNNNNNNN | 1 |
| R | 6118 hCoV-19/Equatorial Guinea/7893/2020\|EPI_ISL_648348 | ···················· | ···················· | ···················· | ···················· | NNNNNNNNNNNNNNNNNNNN | NNNNNNNNN····NN····· | 1 |
| R | 6119 hCoV-19/Equatorial Guinea/8844/2020\|EPI_ISL_648350 | ···················· | ···················· | ···················· | ···················· | NNNNNNNNNNNNNNNNNNNN | NNNNNNNNNNNNNNNNNNNN | 1 |
| R | 6120 hCoV-19/Equatorial Guinea/9134/2020\|EPI_ISL_648352 | ···················· | ···················· | ···················· | ···················· | NNNNNNNNNNNNNNNNNNNN | NNNNNNNNNNNNNNNNNNNN | 1 |

| | | | | | | | | | |
|---|---|---|---|---|---|---|---|---|---|
| R | 6121 | hCoV-19/Equatorial Guinea/9357/2020\|EPI_ISL_648353 | ···················· | ···················· | ···················· | ···················· | NNNNNNNNNNNN········ | ···················· | 1 |
| R | 6123 | hCoV-19/Equatorial Guinea/9516/2020\|EPI_ISL_648355 | ···················· | ···················· | ···················· | ···················· | NNNNNNNNNNNNNNNNNNNN | NNNNNNNNNNNNNNNNNNNN | 1 |
| R | 6124 | hCoV-19/Equatorial Guinea/21591/2020\|EPI_ISL_64836 | ···················· | ·A·················· | ···················· | ···················· | NNNNNNNNNNNNNNNNNNNN | NNNNNNNNNNNNNNNNNNNN | 1 |
| R | 6125 | hCoV-19/Equatorial Guinea/29422/2020\|EPI_ISL_64837 | ···················· | ···················· | ···················· | ···················· | NNNNNNNNNNNNNNNNNNNN | NNNNNNNNNNNNNNNNNNNN | 1 |
| R | 6126 | hCoV-19/Equatorial Guinea/29828/2020\|EPI_ISL_64837 | ···················· | ···················· | ···················· | ···················· | NNNNNNNNNNNNNNNNNNNN | NNNNNNNNNNNNNNNNNNNN | 1 |
| R | 6127 | hCoV-19/Equatorial Guinea/47665/2020\|EPI_ISL_64837 | ···················· | ···················· | ···················· | ···················· | NNNNNNNNNNNNNNNNNNNN | NNNNN··NN··········· | 1 |
| R | 6144 | hCoV-19/USA/CA-CZB-12315/2020\|EPI_ISL_648399\|2020- | ···················· | ···················· | ···················· | NNNNNNNNNNNNNNN····· | ···················· | ···················· | 1 |
| R | 6560 | hCoV-19/Belgium/ULG-10766/2020\|EPI_ISL_644612\|2020 | ···················· | ···················· | ···················· | ···················· | NNNNNNNNNNNNNNNNNNNN | NNNNNNNNNNNNNNNNNNNN | 1 |
| R | 6562 | hCoV-19/Belgium/ULG-10768/2020\|EPI_ISL_644614\|2020 | ···················· | ·A·················· | ···················· | ···················· | NNNNNNNNNNNNNNNNNNNN | NNNNNNNNNNNNNNNNNNNN | 1 |
| R | 6565 | hCoV-19/Belgium/ULG-10771/2020\|EPI_ISL_644617\|2020 | ···················· | ···················· | ···················· | ···················· | NNNNNNNNNNNNNNNNNNNN | NNNNNNNNNNNNNNNNNNNN | 1 |
| R | 6567 | hCoV-19/Belgium/ULG-10774/2020\|EPI_ISL_644620\|2020 | ···················· | ···················· | ···················· | ···················· | NNNNNNNNNNNNNNNNNNNN | NNNNNNNNNNNNNNNNNNNN | 1 |
| R | 6568 | hCoV-19/Belgium/ULG-10775/2020\|EPI_ISL_644621\|2020 | ···················· | ···················· | ···················· | ···················· | NNNNNNNNNNNNNNNNNNNN | NNNNNNNNNNNNNNNNNNNN | 1 |
| R | 6572 | hCoV-19/Belgium/ULG-10780/2020\|EPI_ISL_644626\|2020 | ···················· | ···················· | ···················· | ···················· | NNNNNNNNNNNNNNNNNNNN | NNNNNNNNNNNNNNNNNNNN | 1 |
| R | 6584 | hCoV-19/France/OCC-SC467/2020\|EPI_ISL_644683\|2020- | ···················· | ···················A | ···················· | NNNN·NNNNNNNNNNNNNNN | ···················· | ···················· | 1 |
| R | 6967 | hCoV-19/Australia/VIC16599/2020\|EPI_ISL_640952\|202 | ···················· | ···················· | ···················· | ···················· | NNNNNNNNNNNNNNNNNNNN | NNNNNNNNNNNNNNNNNNNN | 1 |
| R | 6977 | hCoV-19/Australia/VIC16256/2020\|EPI_ISL_640963\|202 | ···················· | ···················· | ···················· | ···················· | NNNNNNNNNNNNNNNNN··· | ···················· | 1 |
| R | 6983 | hCoV-19/Australia/VIC16418/2020\|EPI_ISL_640971\|202 | ···················· | ···················· | ···················· | ···················· | ···················· | NNNNNNNNNNNNNNN····· | 1 |
| R | 6986 | hCoV-19/Australia/VIC16210/2020\|EPI_ISL_640975\|202 | ···················· | ···················· | ···················· | ···················· | NNNNNNNNNNNNNNNNNN·· | ···················· | 1 |
| R | 6988 | hCoV-19/Australia/VIC16277/2020\|EPI_ISL_640977\|202 | ···················· | ···················· | ···················· | ···················· | NNNNNNNNNNNNNNNNNNNN | NNNNNNNNNNNNNNN····· | 1 |
| R | 7200 | hCoV-19/Australia/VIC16652/2020\|EPI_ISL_640914\|202 | ···················· | ···················· | ···················· | ···················· | NNNNNNNNN··········· | ···················· | 1 |
| R | 7203 | hCoV-19/Australia/VIC12530/2020\|EPI_ISL_640917\|202 | ···················· | ·····R·············· | ···················· | ···················· | NNNNNNNNNNNNNNNNNNNN | ···················· | 1 |
| R | 7207 | hCoV-19/Australia/VIC12688/2020\|EPI_ISL_640922\|202 | ···················· | ···················· | ···················· | ···················· | NNNNNNNNNNNNNNNNNNNN | NNNNNNNNNNNNNNN····· | 1 |
| R | 7216 | hCoV-19/Australia/VIC12655/2020\|EPI_ISL_640947\|202 | ···················· | ···················· | ···················· | ···················· | NNNNNNNNNNNNNNNNNNNN | NNNNNNNNNNNNNNNNNNNN | 1 |
| R | 7221 | hCoV-19/Australia/VIC16617/2020\|EPI_ISL_640997\|202 | ···················· | ···················· | ···················· | ···················· | NNNNNNNNNNNNNNNNNNNN | NNNNNNNNNNNNNNNNNNNN | 1 |
| R | 7530 | hCoV-19/France/PAC-MEPHI-1839/2020\|EPI_ISL_644447\| | ···················· | ···················· | ···················· | ···················· | ···················· | NNNNNNNN············ | 1 |
| S | 230 | hCoV-19/Burkina Faso/1689/2020\|EPI_ISL_660448\|2020 | ···················· | ···················· | ···················· | ···················· | NNNNNNNNNNNNNNNNNNNN | NNNNNNNNNNNNNNNNNNNN | 1 |
| S | 256 | hCoV-19/Burkina Faso/583/2020\|EPI_ISL_660474\|2020- | ···················· | ···················· | ···················· | ···················· | NNNNNNNNNNNNNNNNNNNN | NNNNNNNNNNNNNNNNNNNN | 1 |
| S | 262 | hCoV-19/USA/FL-BPHL-2058/2020\|EPI_ISL_653211\|2020- | ···················· | ···················· | ···················· | ···················· | NNNN················ | ···················· | 1 |
| S | 269 | hCoV-19/USA/FL-BPHL-2075/2020\|EPI_ISL_653228\|2020- | ···················· | ···················· | ···················· | ···················· | NNNNNNNNNNNNNNNNNNNN | ···················· | 1 |
| S | 270 | hCoV-19/USA/FL-BPHL-2079/2020\|EPI_ISL_653232\|2020- | ···················· | ···················· | ···················· | ···················· | NNNNNNNNNNNNNNNNNNNN | NNNNNNNNNNNNNNNNNNNN | 1 |
| S | 272 | hCoV-19/USA/FL-BPHL-2083/2020\|EPI_ISL_653236\|2020- | ···················· | ···················· | ···················· | ···················· | NNNNNNNNNNNNNNNNNNNN | NNNNNNNNNNNNNNNNNNNN | 1 |
| S | 296 | hCoV-19/USA/FL-BPHL-2128/2020\|EPI_ISL_653281\|2020- | ···················· | ···················· | ···················· | ···················· | NNNNNNNNNNNNNNNNNNNN | ···················· | 1 |
| S | 302 | hCoV-19/Spain/CT-IBV-97007949/2020\|EPI_ISL_654516\| | ···················· | ···················· | ···················· | ···················· | NNNNNNNNNNNNNNNNNNNN | NNNNNNNNNNNNNNNNNNNN | 1 |
| S | 303 | hCoV-19/Spain/CT-IBV-97007954/2020\|EPI_ISL_654517\| | ···················· | ···················· | ···················· | ···················· | NNNNNNNNNNNNNNNNNNNN | NNNNNNNNNNNNNNNNNNNN | 1 |
| S | 304 | hCoV-19/Spain/CT-IBV-97007969/2020\|EPI_ISL_654518\| | ···················· | ···················· | ···················· | ···················· | NNNNNNNNNNNNNNNNNNNN | NNNNNNNNNNNNNNNNNNNN | 1 |
| S | 305 | hCoV-19/Spain/CT-IBV-97007975/2020\|EPI_ISL_654519\| | ···················· | ···················· | ···················· | ···················· | NNNNNNNNNNNNNNNNNNNN | NNNNNNNNNNNNNNNNNNNN | 1 |
| S | 306 | hCoV-19/Spain/CT-IBV-97007979/2020\|EPI_ISL_654520\| | ···················· | ···················· | ···················· | ···················· | NNNNNNNNNNNNNNNNNNNN | NNNNNNNNNNNNNNNNNNNN | 1 |
| S | 307 | hCoV-19/Spain/CT-IBV-97007980/2020\|EPI_ISL_654521\| | ···················· | ···················· | ···················· | ···················· | NNNNNNNNNNNNNNNNNNNN | NNNNNNNNNNNNNNNNNNNN | 1 |
| S | 308 | hCoV-19/Spain/CT-IBV-97007987/2020\|EPI_ISL_654522\| | ···················· | ···················· | ···················· | ···················· | NNNNNNNNNNNNNNNNNNNN | NNNNNNNNNNNNNNNNNNNN | 1 |
| S | 309 | hCoV-19/Spain/CT-IBV-97007956/2020\|EPI_ISL_654523\| | ···················· | ···················· | ···················· | ···················· | NNNNNNN············· | ···················· | 1 |
| S | 310 | hCoV-19/Spain/CT-IBV-97007960/2020\|EPI_ISL_654524\| | ···················· | ···················· | ···················· | ···················· | NNNNNNNNNNNNNNNNNNNN | NNNNNNNNNNNNNNNNNNNN | 1 |
| S | 312 | hCoV-19/Spain/CT-IBV-97007970/2020\|EPI_ISL_654526\| | ···················· | ···················· | ···················· | ···················· | NNNNNNNNNNNNNNNNNNNN | NNNNNNNNNNNNNNNNNNNN | 1 |
| S | 313 | hCoV-19/Spain/CT-IBV-97007968/2020\|EPI_ISL_654527\| | ···················· | ···················· | ···················· | ···················· | NNNNNNN············· | NNNNNNN······NNNNNNN | 1 |
| S | 314 | hCoV-19/Spain/CT-IBV-97007981/2020\|EPI_ISL_654528\| | ···················· | ···················· | ···················· | ···················· | NNNNNNNNNNNNNNNNNNNN | NNNNNNNNNNNNNNNNNNNN | 1 |
| S | 315 | hCoV-19/Spain/CT-IBV-97007955/2020\|EPI_ISL_654529\| | ···················· | ···················· | ···················· | ···················· | NNNNNNNNNNNNNNNNNNNN | NNNNNNNNNNNNNNNNNNNN | 1 |
| S | 316 | hCoV-19/Spain/CT-IBV-97007944/2020\|EPI_ISL_654530\| | ···················· | ···················· | ···················· | ···················· | NNNNNNNNNNNNNNNNNNNN | NNNNNNNNNNNNNNNNNNNN | 1 |
| S | 317 | hCoV-19/Spain/CT-IBV-97007951/2020\|EPI_ISL_654531\| | ···················· | ···················· | ···················· | ···················· | ·NNNNNNNNNNNNNN····· | ···················· | 1 |
| S | 318 | hCoV-19/Spain/CT-IBV-97007950/2020\|EPI_ISL_654532\| | ···················· | ···················· | ···················· | ···················· | ·NNNNNN············· | ···················· | 1 |
| S | 319 | hCoV-19/Spain/CT-IBV-97007961/2020\|EPI_ISL_654533\| | ···················· | ···················· | ···················· | ···················· | NNNNNNNNNNNNNNNNNNNN | NNNNNNNNNNNNNNNNNNNN | 1 |
| S | 320 | hCoV-19/Spain/CT-IBV-97007971/2020\|EPI_ISL_654534\| | ···················· | ···················· | ···················· | ···················· | NNNNNNNNNNNNNNNNNNNN | NNNNNNNNNNNNNNNNNNNN | 1 |
| S | 321 | hCoV-19/Spain/CT-IBV-97007984/2020\|EPI_ISL_654535\| | ···················· | ···················· | ···················· | ···················· | NNNNNNNNNNNNNNNNNNNN | NNNNNNNNNNNNNNNNNNNN | 1 |
| S | 322 | hCoV-19/Spain/CT-IBV-97007989/2020\|EPI_ISL_654536\| | ···················· | ···················· | ···················· | ···················· | NNNNNNN············· | ···················· | 1 |
| S | 323 | hCoV-19/Spain/CT-IBV-97007973/2020\|EPI_ISL_654537\| | ···················· | ···················· | ···················· | ···················· | NNNNNNNNNNNNNNNNNNNN | NNNNNNNNNNNNNNNNNNNN | 1 |
| S | 324 | hCoV-19/Spain/CT-IBV-97007953/2020\|EPI_ISL_654538\| | ···················· | ···················· | ···················· | ···················· | NNNNNNNNNNNNNNNNNNNN | NNNNNNNNNNNNNNNNNNNN | 1 |
| S | 496 | hCoV-19/Burkina Faso/2-032-1197/2020\|EPI_ISL_66047 | ···················· | ···················· | ···················· | ···················· | NNNNNNNNNNNNNNNNNNNN | NNNNNNNNNNNNNNNNNNNN | 1 |
| S | 502 | hCoV-19/Burkina Faso/2453/2020\|EPI_ISL_660485\|2020 | ···················· | ···················· | ···················· | ···················· | NNNNNNNNNNNNNNNNNNNN | NNNNNNNNNNNNNNNNNNNN | 1 |
| S | 517 | hCoV-19/USA/MI-UM-10036596238/2020\|EPI_ISL_649978\| | ···················· | ···················· | ···················· | ···················· | NNNNNNNNNNNNNNNNNNNN | ···················· | 1 |
| S | 519 | hCoV-19/USA/MI-UM-10036578395/2020\|EPI_ISL_649981\| | ···················· | ···················· | ···················· | ···················· | NNNNNNNNNNNNNNNNNNNN | NNNNNNNNNNNNNNNNNNNN | 1 |
| S | 521 | hCoV-19/USA/MI-UM-10036598438/2020\|EPI_ISL_649983\| | ···················· | ···················· | ···················· | ···················· | NNNNNNNNNNNNNNNNNNNN | ···················· | 1 |
| S | 522 | hCoV-19/USA/MI-UM-10036587493/2020\|EPI_ISL_649984\| | ···················· | ···················· | ···················· | ···················· | NNNNNNNNNNNNNNNNNNNN | ···················· | 1 |
| S | 525 | hCoV-19/USA/MI-UM-10036587385/2020\|EPI_ISL_649988\| | ···················· | ···················· | ···················· | ···················· | NNNNNNNNNNNNNNNNNNNN | ···················· | 1 |
| S | 526 | hCoV-19/USA/MI-UM-10036587181/2020\|EPI_ISL_649989\| | ···················· | ···················· | ···················· | ···················· | NNNNNNNNNNNNNNNNNNNN | ···················· | 1 |
| S | 527 | hCoV-19/USA/MI-UM-10036598672/2020\|EPI_ISL_649990\| | ···················· | ···················· | ···················· | ···················· | NNNNNNNNNNNNNNNNNNNN | ···················· | 1 |
| S | 529 | hCoV-19/USA/MI-UM-10036598494/2020\|EPI_ISL_649992\| | ···················· | ···················· | ···················· | ···················· | NNNNNNNNNNNNNNNNNNNN | ···················· | 1 |
| S | 690 | hCoV-19/Spain/CT-IBV-99009903/2020\|EPI_ISL_660300\| | ···················· | ···················· | ···················· | ···················· | NNNNNNNNNNNNNNNNNNNN | NNNNNNNNNNNNNNNNNNNN | 1 |
| S | 691 | hCoV-19/Spain/CT-IBV-99009904/2020\|EPI_ISL_660301\| | ···················· | ···················· | ···················· | ···················· | NNNNNNNNNNNNNNNNNNNN | NNNNNNNNNNNNNNNNNNNN | 1 |
| S | 692 | hCoV-19/Spain/CT-IBV-99009905/2020\|EPI_ISL_660302\| | ···················· | ···················· | ···················· | ···················· | ··NNNNN············· | ···················· | 1 |
| S | 693 | hCoV-19/Spain/CT-IBV-99009906/2020\|EPI_ISL_660303\| | ···················· | ···················· | ···················· | ···················· | NNNNNNNNNNNNNNNNNNNN | NNNNNNNNNNNNNNNNNNNN | 1 |
| S | 694 | hCoV-19/Spain/CT-IBV-99009907/2020\|EPI_ISL_660304\| | ···················· | ···················· | ···················· | ···················· | NNNNNNNNNNNNNNNNNNNN | NNNNNNNNNNNNNNNNNNNN | 1 |
| S | 695 | hCoV-19/Spain/CT-IBV-99009908/2020\|EPI_ISL_660305\| | ···················· | ···················· | ···················· | ···················· | ··NNNNN··NNNNNNNNNNN | ···················· | 1 |
| S | 696 | hCoV-19/Spain/CT-IBV-99009909/2020\|EPI_ISL_660306\| | ···················· | ···················· | ···················· | ···················· | NNNNNNNNNNNNNNNNNNNN | NNNNNNNNNNNNNNNNNNNN | 1 |
| S | 697 | hCoV-19/Spain/CT-IBV-99009910/2020\|EPI_ISL_660307\| | ···················· | ···················· | ···················· | ···················· | ··NNNNNNNNNNNNNNNNNN | ···················· | 1 |
| S | 698 | hCoV-19/Spain/CT-IBV-99009911/2020\|EPI_ISL_660308\| | ···················· | ···················· | ···················· | ···················· | NNNNNNNNNNNNNNNNNNNN | NNNNNNNNNNNNNNNNNNNN | 1 |
| S | 699 | hCoV-19/Spain/CT-IBV-99009912/2020\|EPI_ISL_660309\| | ···················· | ···················· | ···················· | ···················· | NNNNNNNNNNNNNNNNNNNN | NNNNNNNNNNNNNNNNNNNN | 1 |
| S | 700 | hCoV-19/Spain/CT-IBV-99009913/2020\|EPI_ISL_660310\| | ···················· | ···················· | ···················· | ···················· | NNNNNNNNNNNNNNNNNNNN | NNNNNNNNNNNNNNNNNNNN | 1 |
| S | 701 | hCoV-19/Spain/CT-IBV-99009914/2020\|EPI_ISL_660311\| | ···················· | ···················· | ···················· | ···················· | NNNNNNNNNNNNNNNNNNNN | NNNNNNNNNNNNNNNNNNNN | 1 |
| S | 702 | hCoV-19/Spain/CT-IBV-99009915/2020\|EPI_ISL_660312\| | ···················· | ···················· | ···················· | ···················· | NNNNNNNNNNNNNNNNNNNN | ···················· | 1 |
| S | 703 | hCoV-19/Spain/CT-IBV-99009916/2020\|EPI_ISL_660313\| | ···················· | ···················· | ···················· | ···················· | ··NNNNN············· | ···················· | 1 |
| S | 705 | hCoV-19/Spain/CT-IBV-99009918/2020\|EPI_ISL_660315\| | ···················· | ···················· | ···················· | ···················· | ·NNNNNNNNNNNNNNNNNNN | ·N·················· | 1 |
| S | 706 | hCoV-19/Spain/CT-IBV-99009919/2020\|EPI_ISL_660316\| | ···················· | ···················· | ···················· | ···················· | NNNNNNNNNNNNNNNNNNNN | NNNNNNNNNNNNNNNNNNNN | 1 |

| | | | | | | | | |
|---|---|---|---|---|---|---|---|---|
| S | 707 hCoV-19/Spain/CT-IBV-99009922/2020\|EPI_ISL_660318\| | ···················· | ···················· | ···················· | ···················· | ·NNNNNNNNNNNNNNNNNNN | NNNNNNNNNNNNNNNNNNNN | 1 |
| S | 708 hCoV-19/Spain/CT-IBV-99009923/2020\|EPI_ISL_660319\| | ···················· | ···················· | ···················· | ···················· | ··NNNNN············· | ·····NNNNN·········· | 1 |
| S | 709 hCoV-19/Spain/CT-IBV-99009924/2020\|EPI_ISL_660320\| | ···················· | ···················· | ···················· | ···················· | ·NNNNNNNNNNNNNNNNNNN | NNNNNNNNNNNNNNNNNNNN | 1 |
| S | 710 hCoV-19/Spain/CT-IBV-99009925/2020\|EPI_ISL_660321\| | ···················· | ···················· | ···················· | ···················· | NNNNNNNNNNNNNNNNNNNN | NNNNNNNNNNNNNNNNNNNN | 1 |
| S | 711 hCoV-19/Spain/CT-IBV-99009926/2020\|EPI_ISL_660322\| | ···················· | ···················· | ···················· | ···················· | NNNNNNNNNNNNNNNNNNNN | NNNNNNNNNNNNNNNNNNNN | 1 |
| S | 813 hCoV-19/USA/FL-BPHL-1955/2020\|EPI_ISL_653108\|2020- | ···················· | ···················· | ···················· | ···················· | NNNNNNNNNNNNNNNNNNNN | ···················· | 1 |
| S | 830 hCoV-19/USA/FL-BPHL-1975/2020\|EPI_ISL_653128\|2020- | ···················· | ···················· | ···················· | ···················· | NNNNNNNNNNNNNNNNNNNN | NNNNNNNNNNNNNNNNNNNN | 1 |
| S | 843 hCoV-19/USA/FL-BPHL-2004/2020\|EPI_ISL_653157\|2020- | ···················· | ···················· | ···················· | ···················· | NNNNNNNNNNNNNNNNNNNN | NNNNNNNNNNNNNNNNNNNN | 1 |
| S | 845 hCoV-19/USA/FL-BPHL-2010/2020\|EPI_ISL_653163\|2020- | ···················· | ···················· | ···················· | ···················· | NNNNNNNNNNNNNNNNNNNN | ···················· | 1 |
| S | 883 hCoV-19/Spain/AR-IBV-98008125/2020\|EPI_ISL_654458\| | ···················· | ···················· | ···················· | ···················· | ··NNNNNNNNNNNNNNNNNN | ···················· | 1 |
| S | 892 hCoV-19/Spain/AR-IBV-98008166/2020\|EPI_ISL_654467\| | ···················· | ···················· | ···················· | ···················· | NNNNNNNNNNNNNNNNNNNN | NNNNNNNNNNNNNNN····· | 1 |
| S | 1013 hCoV-19/South Africa/KRISP-K004357-CE1/2020\|EPI_IS | ···················· | ···················· | ···················· | ···················· | NNNNNNNNNNNNNNNNN··· | ···················· | 1 |
| S | 1087 hCoV-19/Spain/MD-IBV-99008780/2020\|EPI_ISL_654323\| | ···················· | ···················· | ···················· | ···················· | NNNNNNNNNNNNN······· | N··················· | 1 |
| S | 1091 hCoV-19/Spain/MD-IBV-99009629/2020\|EPI_ISL_654338\| | ···················· | ···················· | ···················· | ···················· | NNNNNNNNNNNNNNNNNNNN | NNNNNNNNNNNNNNNNNNNN | 1 |
| S | 1092 hCoV-19/Spain/MD-IBV-99009630/2020\|EPI_ISL_654339\| | ···················· | ···················· | ···················· | ···················· | NNNNNNNNNNNNNNNNNNNN | NNNNNNNNNNNNNNNNNNNN | 1 |
| S | 1093 hCoV-19/Spain/MD-IBV-99009631/2020\|EPI_ISL_654340\| | ···················· | ···················· | ···················· | ···················· | NNNNNNNNNNNNNNNNNNNN | NNNNNNNNNNNNNNNNNNNN | 1 |
| S | 1094 hCoV-19/Spain/MD-IBV-99009663/2020\|EPI_ISL_654341\| | ···················· | ···················· | ···················· | ···················· | NNNNNNNNNNNNNNNNNNNN | NNNNNNNNNNNNNNNNNNNN | 1 |
| S | 1097 hCoV-19/Spain/MD-IBV-99009592/2020\|EPI_ISL_654344\| | ···················· | ···················· | ···················· | ···················· | NNNNNNNNNNNNNNNNNNNN | ············NNN····· | 1 |
| S | 1101 hCoV-19/Spain/MD-IBV-99009596/2020\|EPI_ISL_654348\| | ···················· | ···················· | ···················· | ···················· | NNNNNNN·········NNNN | ···················· | 1 |
| S | 1102 hCoV-19/Spain/MD-IBV-99009597/2020\|EPI_ISL_654349\| | ···················· | ···················· | ···················· | ···················· | NNNNNNNNNNNNNNNNNNNN | NNNNNNNNNNNNNNNNNNNN | 1 |
| S | 1103 hCoV-19/Spain/MD-IBV-99009601/2020\|EPI_ISL_654350\| | ···················· | ···················· | ···················· | ···················· | NNNNNNNNNNNNNNNNNNNN | NNNNNNN···········NN | 1 |
| S | 1104 hCoV-19/Spain/MD-IBV-99009602/2020\|EPI_ISL_654351\| | ···················· | ···················· | ···················· | ···················· | NNNNNNNNNNNNNNNNNNNN | NNNNNNNNNNNNNNNNNNNN | 1 |
| S | 1105 hCoV-19/Spain/MD-IBV-99009603/2020\|EPI_ISL_654352\| | ···················· | ···················· | ···················· | ···················· | NNNNNNN············· | NNNNNNNNNNNNNNN····· | 1 |
| S | 1106 hCoV-19/Spain/MD-IBV-99009604/2020\|EPI_ISL_654353\| | ···················· | ···················· | ···················· | ···················· | NNNNNNNNNNNNNNNNNNNN | NNNNNNNNNNNNNNNNNNNN | 1 |
| S | 1108 hCoV-19/Spain/MD-IBV-99009606/2020\|EPI_ISL_654355\| | ···················· | ···················· | ···················· | ···················· | NNNNNNNNNNNNNNNNNNNN | NNNNNNNNNNNNNNNNNNNN | 1 |
| S | 1109 hCoV-19/Spain/MD-IBV-99009607/2020\|EPI_ISL_654356\| | ···················· | ···················· | ···················· | ···················· | NNNNNNNNNNNNNNNNNNNN | NNNNNNNNNNNNNNNNNNNN | 1 |
| S | 1110 hCoV-19/Spain/MD-IBV-99009608/2020\|EPI_ISL_654357\| | ···················· | ···················· | ···················· | ···················· | NNNNNNNNNNNNNNNNNNNN | NNNNNNNNNNNNNNNNNNNN | 1 |
| S | 1111 hCoV-19/Spain/MD-IBV-99009609/2020\|EPI_ISL_654358\| | ···················· | ···················· | ···················· | ···················· | NNNNNNNNNNNNNNNNNNNN | NNNNNNNNNNNNNNNNNNNN | 1 |
| S | 1247 hCoV-19/Spain/CT-IBV-99009869/2020\|EPI_ISL_660266\| | ···················· | ···················· | ···················· | ···················· | ·NNNNNN···········NN | ···················· | 1 |
| S | 1256 hCoV-19/Spain/CT-IBV-99009878/2020\|EPI_ISL_660275\| | ···················· | ···················· | ···················· | ···················· | NNNNNNNNNNNNNNNNNNNN | ···················· | 1 |
| S | 1257 hCoV-19/Spain/CT-IBV-99009879/2020\|EPI_ISL_660276\| | ···················· | ···················· | ···················· | ···················· | ··NNNNN············· | ···················· | 1 |
| S | 1264 hCoV-19/Spain/CT-IBV-99009886/2020\|EPI_ISL_660283\| | ···················· | ···················· | ···················· | ···················· | NNNNNNN············· | ···················· | 1 |
| S | 1269 hCoV-19/Spain/CT-IBV-99009891/2020\|EPI_ISL_660288\| | ···················· | ···················· | ···················· | ···················· | ··NNNNN············· | NNNNNNNNNNNNNNN····· | 1 |
| S | 1271 hCoV-19/Spain/CT-IBV-99009893/2020\|EPI_ISL_660290\| | ···················· | ···················· | ···················· | ···················· | NNNNNNNNNNNNNNNNNNNN | NNNNNNNNNNNNNNNNNNNN | 1 |
| S | 1277 hCoV-19/Spain/CT-IBV-99009899/2020\|EPI_ISL_660296\| | ···················· | ···················· | ···················· | ···················· | NNNNNNN············· | ···················· | 1 |
| S | 1278 hCoV-19/Spain/CT-IBV-99009900/2020\|EPI_ISL_660297\| | ···················· | ···················· | ···················· | ···················· | NNNNNNNNNNNNNNNNNNNN | NNNNNNNNNNNNNNNNNNNN | 1 |
| S | 1279 hCoV-19/Spain/CT-IBV-99009901/2020\|EPI_ISL_660298\| | ···················· | ···················· | ···················· | ···················· | NNNNNNNNNNNNNNNNNNNN | NNNNNNNNNNNNNNNNNNNN | 1 |
| S | 1280 hCoV-19/Spain/CT-IBV-99009902/2020\|EPI_ISL_660299\| | ···················· | ···················· | ···················· | ···················· | NNNNNNNNNNNNNNNNNNNN | NNNNNNNNNNNNNNNNNNNN | 1 |
| S | 1382 hCoV-19/Spain/MD-IBV-99008726/2020\|EPI_ISL_654300\| | ···················· | ···················· | ···················· | ···················· | NNNNNNNNNNNNNNNNNNNN | NNNNNNNNNNNNNNNNNNNN | 1 |
| S | 1384 hCoV-19/Spain/MD-IBV-99008729/2020\|EPI_ISL_654302\| | ···················· | ···················· | ···················· | ···················· | NNNNNNNNNNNNNNNNNNNN | NNNNNNNNNNNNNNNNNNNN | 1 |
| S | 1385 hCoV-19/Spain/MD-IBV-99008734/2020\|EPI_ISL_654303\| | ···················· | ···················· | ···················· | ···················· | ··NNNNN··NNNNNNNNNNN | NNNNNNNNNNNNNNNNNNNN | 1 |
| S | 1386 hCoV-19/Spain/MD-IBV-99008736/2020\|EPI_ISL_654304\| | ···················· | ···················· | ···················· | ···················· | NNNNNNNNNNNNNNNNNNNN | NNNNNNNNNNNNNNNNNNNN | 1 |
| S | 1387 hCoV-19/Spain/MD-IBV-99008738/2020\|EPI_ISL_654305\| | ···················· | ···················· | ···················· | ···················· | NNNNNNNNNNNNNNNNNNNN | NNNNNNNNNNNNNNNNNNNN | 1 |
| S | 1388 hCoV-19/Spain/MD-IBV-99008739/2020\|EPI_ISL_654306\| | ···················· | ···················· | ···················· | ···················· | NNNNNNNNNNNNNNNNNNNN | NNNNNNNNNNNNNNNNNNNN | 1 |
| S | 1389 hCoV-19/Spain/MD-IBV-99008741/2020\|EPI_ISL_654307\| | ···················· | ···················· | ···················· | ···················· | NNNNNNNNNNNNNNNNNNNN | NNNNNNNNNNNNNNNNNNNN | 1 |
| S | 1390 hCoV-19/Spain/MD-IBV-99008744/2020\|EPI_ISL_654308\| | ···················· | ···················· | ···················· | ···················· | NNNNNNNNNNNNNNNNNNNN | NNNNNNNNNNNNNNNNNNNN | 1 |
| S | 1391 hCoV-19/Spain/MD-IBV-99008747/2020\|EPI_ISL_654309\| | ···················· | ···················· | ···················· | ···················· | NNNNNNNNNNNNNNNNNNNN | NNNNNNNNNNNNNNNNNNNN | 1 |
| S | 1392 hCoV-19/Spain/MD-IBV-99008748/2020\|EPI_ISL_654310\| | ···················· | ···················· | ···················· | ···················· | NNNNNNNNNNNNNNNNNNNN | NNNNNNNNNNNNNNNNNNNN | 1 |
| S | 1394 hCoV-19/Spain/MD-IBV-99008754/2020\|EPI_ISL_654312\| | ···················· | ···················· | ···················· | ···················· | NNNNNNNNNNNNNNNNNNNN | NNNNNNNNNNNNNNNNNNNN | 1 |
| S | 1395 hCoV-19/Spain/MD-IBV-99008755/2020\|EPI_ISL_654313\| | ···················· | ···················· | ···················· | ···················· | NNNNNNNNNNNNNNNNNNNN | NNNNNNNNNNNNNNNNNNNN | 1 |
| S | 1396 hCoV-19/Spain/MD-IBV-99008757/2020\|EPI_ISL_654314\| | ···················· | ···················· | ···················· | ···················· | NNNNNNNNNNNNNNNNNNNN | NNNNNNNNNNNNNNNNNNNN | 1 |
| S | 1397 hCoV-19/Spain/MD-IBV-99008758/2020\|EPI_ISL_654315\| | ···················· | ···················· | ···················· | ···················· | NNNNNNNNNNNNNNNNNNNN | NNNNNNNNNNNNNNNNNNNN | 1 |
| S | 1398 hCoV-19/Spain/MD-IBV-99008761/2020\|EPI_ISL_654316\| | ···················· | ···················· | ···················· | ···················· | NNNNNNNNNNNNNNNNNNNN | NNNNNNNNNNNNNNNNNNNN | 1 |
| S | 1399 hCoV-19/Spain/MD-IBV-99008762/2020\|EPI_ISL_654317\| | ···················· | ···················· | ···················· | ···················· | NNNNNNNNNNNNNNNNNNNN | NNNNNNNNNNNNNNNNNNNN | 1 |
| S | 1400 hCoV-19/Spain/MD-IBV-99008765/2020\|EPI_ISL_654318\| | ···················· | ···················· | ···················· | ···················· | NNNNNNNNNNNNNNNNNNNN | NNNNNNNNNNNNNNNNNNNN | 1 |
| S | 1402 hCoV-19/Spain/MD-IBV-99008779/2020\|EPI_ISL_654322\| | ···················· | ···················· | ···················· | ···················· | NNNN················ | ···················· | 1 |
| S | 1403 hCoV-19/Spain/MD-IBV-99008783/2020\|EPI_ISL_654326\| | ···················· | ···················· | ···················· | ···················· | NNNNNNNNNNNNNNNNNNNN | ··············NNNNNN | 1 |
| S | 1404 hCoV-19/Spain/MD-IBV-99008787/2020\|EPI_ISL_654327\| | ···················· | ···················· | ···················· | ···················· | NNNNNNNNNNNNNNNNNNNN | NNNNNNNNNNNNNNNNNNNN | 1 |
| S | 1406 hCoV-19/Spain/MD-IBV-99009611/2020\|EPI_ISL_654360\| | ···················· | ···················· | ···················· | ···················· | NNNNNNNNNNNNNNNNNNNN | NNNNNNNNNNNNNNNNNNNN | 1 |
| S | 1407 hCoV-19/Spain/MD-IBV-99009615/2020\|EPI_ISL_654361\| | ···················· | ···················· | ···················· | ···················· | NNNNNNNNNNNNNNNNNNNN | NNNNNNNNNNNNNNNNNNNN | 1 |
| S | 1409 hCoV-19/Spain/MD-IBV-99009622/2020\|EPI_ISL_654363\| | ···················· | ···················· | ···················· | ···················· | NNNNNNNNNNNNNNNNNNNN | NNNNNNNNNNNNNNNNNNNN | 1 |
| S | 1410 hCoV-19/Spain/MD-IBV-99009623/2020\|EPI_ISL_654364\| | ···················· | ···················· | ···················· | ···················· | NNNNNNNNNNNNNNNNNNNN | NNNNNNNNNNNNNNNNNNNN | 1 |
| S | 1411 hCoV-19/Spain/MD-IBV-99009624/2020\|EPI_ISL_654365\| | ···················· | ···················· | ···················· | ···················· | NNNNNNNNNNNNNNNNNNNN | NNNNNNNNNNNNNNNNNNNN | 1 |
| S | 1413 hCoV-19/Spain/MD-IBV-99009632/2020\|EPI_ISL_654369\| | ···················· | ···················· | ···················· | ···················· | NNNNNNNNNNNNNNNNNNNN | NNNNNNNNNNNNNNNNNNNN | 1 |
| S | 1414 hCoV-19/Spain/MD-IBV-99009645/2020\|EPI_ISL_654380\| | ···················· | ···················· | ···················· | ···················· | NNNNNNNNNNNNNNNNNNNN | NNNNNNNNNNNNNNNNNNNN | 1 |
| S | 1415 hCoV-19/Spain/MD-IBV-99009647/2020\|EPI_ISL_654381\| | ···················· | ···················· | ···················· | ···················· | NNNNNNNNNNNNNNNNNNNN | NNNNNNNNNNNNNNNNNNNN | 1 |
| S | 1417 hCoV-19/Spain/MD-IBV-99009649/2020\|EPI_ISL_654383\| | ···················· | ···················· | ···················· | ···················· | NNNNNNNNNNNNNNNNNNNN | NNNNNNNNNNNNNNNNNNNN | 1 |
| S | 1418 hCoV-19/Spain/MD-IBV-99009654/2020\|EPI_ISL_654384\| | ···················· | ···················· | ···················· | ···················· | NNNNNNNNNNNNNNNNNNNN | NNNNNNNNNNNNNNNNNNNN | 1 |
| S | 1419 hCoV-19/Spain/MD-IBV-99009655/2020\|EPI_ISL_654385\| | ······C············· | ···················· | ···················· | ···················· | NNNNNNNNNNNNNNNNNNNN | NNNNNNNNNNNNNNNNNNNN | 1 |
| S | 1420 hCoV-19/Spain/MD-IBV-99009656/2020\|EPI_ISL_654386\| | ···················· | ···················· | ···················· | ···················· | NNNNNNNNNNNNNNNNNNNN | ··············N··NNN | 1 |
| S | 1421 hCoV-19/Spain/MD-IBV-99009657/2020\|EPI_ISL_654387\| | ···················· | ···················· | ···················· | ···················· | NNNNNNNNNNNNNNNNNNNN | NNNNNNNNNNNNNNNNNNNN | 1 |
| S | 1422 hCoV-19/Spain/MD-IBV-99009660/2020\|EPI_ISL_654388\| | ···················· | ···················· | ···················· | ···················· | NNNNNNNNNNNNNNNNNNNN | NNNNNNNNNNNNNNNNNNNN | 1 |
| S | 1423 hCoV-19/Spain/MD-IBV-99009661/2020\|EPI_ISL_654389\| | ···················· | ···················· | ···················· | ···················· | NNNNNNNNNNNNNNNNNNNN | NNNNNNNNNNNNNNNNNNNN | 1 |
| S | 1424 hCoV-19/Spain/MD-IBV-99009662/2020\|EPI_ISL_654390\| | ···················· | ···················· | ···················· | ···················· | NNNNNNNNNNNNNNNNNNNN | ···················· | 1 |
| S | 1425 hCoV-19/Spain/MD-IBV-99009664/2020\|EPI_ISL_654391\| | ···················· | ···················· | ···················· | ···················· | ·NNNN··············· | ·····NNNNNNNNNN····· | 1 |
| S | 1426 hCoV-19/Spain/MD-IBV-99009667/2020\|EPI_ISL_654392\| | ···················· | ···················· | ···················· | ···················· | NNNNNNNNNNNNNNNNNNNN | NNNNNNNNNNNNNNNNNNNN | 1 |
| S | 1427 hCoV-19/Spain/MD-IBV-99009668/2020\|EPI_ISL_654393\| | ···················· | ···················· | ···················· | ···················· | NNNNNNNNNNNNNNNNNNNN | NNNNNNNNNNNNNNNNN··· | 1 |
| S | 1428 hCoV-19/Spain/MD-IBV-99009669/2020\|EPI_ISL_654394\| | ···················· | ···················· | ···················· | ···················· | NNNNNNNNNNNNNNNNNNNN | NNNNNNNNNNNNNNNNNNNN | 1 |
| S | 1429 hCoV-19/Spain/MD-IBV-99009670/2020\|EPI_ISL_654395\| | ···················· | ···················· | ···················· | ···················· | NNNNNNNNNNNNNNNNNNNN | NNNNNNNNNNNNNNNNNNNN | 1 |
| S | 1430 hCoV-19/Spain/MD-IBV-99009671/2020\|EPI_ISL_654396\| | ···················· | ···················· | ···················· | ···················· | NNNNNNNNNNNNNNNNNNNN | NNNNNNNNNNNNNNNNNNNN | 1 |

```
S    1431 hCoV-19/Spain/MD-IBV-99009672/2020|EPI_ISL_654397|    ····················  ····················  ····················  ····················  NNNNNNNNNNNNNNNNNNNN  NNNNNNNNNNNNNNNNNNNN    1
S    1615 hCoV-19/Spain/MD-IBV-99008658/2020|EPI_ISL_654220|    ····················  ····················  ····················  ····················  NNNNNNNNNNNN·NNNN···  ····················    1
S    1618 hCoV-19/Spain/MD-IBV-99008664/2020|EPI_ISL_654225|    ····················  ····················  ····················  ····················  ·NNNNNNNNNNNNNNNNNNN  NNNNNNNNNNNNNNNNNNNN    1
S    1619 hCoV-19/Spain/MD-IBV-99008665/2020|EPI_ISL_654226|    ····················  ····················  ····················  ····················  NNNNNNNNNNNNNNNNNNNN  NNNNNNNNNNNNNNNNNNNN    1
S    1620 hCoV-19/Spain/MD-IBV-99009610/2020|EPI_ISL_654359|    ····················  ····················  ····················  ····················  NNNNNNNNNNNNNNNNNNNN  NNNNNNNNNNNNNNNNNNNN    1
S    1621 hCoV-19/Spain/MD-IBV-99009626/2020|EPI_ISL_654366|    ····················  ····················  ····················  ····················  NNNNNNNNNNNNNNNNNNNN  NNNNNNNNNNNNNNNNNNNN    1
S    1622 hCoV-19/Spain/MD-IBV-99009627/2020|EPI_ISL_654367|    ····················  ····················  ····················  ····················  NNNNNNNNNNNNNNNNNNNN  NNNNNNNNNNNNNNN·NNNN    1
S    1623 hCoV-19/Spain/MD-IBV-99009633/2020|EPI_ISL_654370|    ····················  ····················  ····················  ····················  NNNNNNNNNNNNNNNNNNNN  NNNNNNNNNNNNNNNNNNNN    1
S    1624 hCoV-19/Spain/MD-IBV-99009635/2020|EPI_ISL_654371|    ····················  ····················  ····················  ····················  NNNNNNNNNNNNNNNNNNNN  NNNNNNNNNNNNNNNNNNNN    1
S    1625 hCoV-19/Spain/MD-IBV-99009636/2020|EPI_ISL_654372|    ····················  ····················  ····················  ····················  NNNNNNNNNNNNNNNNNNNN  NNNNNNNNNNNNNNNNNNNN    1
S    1626 hCoV-19/Spain/MD-IBV-99009637/2020|EPI_ISL_654373|    ····················  ····················  ····················  ····················  NNNNNNNNNNNNNNNNNNNN  NNNNNNNNNNNNNNNNNNNN    1
S    1627 hCoV-19/Spain/MD-IBV-99009638/2020|EPI_ISL_654374|    ····················  ····················  ····················  ····················  NNNNNNNNNNNNNNNNNNNN  NNNNNNNNNNNNNNNNNNNN    1
S    1628 hCoV-19/Spain/MD-IBV-99009639/2020|EPI_ISL_654375|    ····················  ····················  ····················  ····················  NNNNNNNNNNNNNNNNNNNN  NNNNNNNNNNNNNNNNNNNN    1
S    1629 hCoV-19/Spain/MD-IBV-99009640/2020|EPI_ISL_654376|    ····················  ····················  ····················  ····················  NNNNNNNNNNNNNNNNNNNN  NNNNNNNNNNNNNNNNNNNN    1
S    1630 hCoV-19/Spain/MD-IBV-99009641/2020|EPI_ISL_654377|    ····················  ····················  ····················  ····················  NNNNNNNNNNNNNNNNNNNN  NNNNNNNNNNNNNNNNNNNN    1
S    1631 hCoV-19/Spain/MD-IBV-99009643/2020|EPI_ISL_654378|    ····················  ····················  ····················  ····················  NNNNNNNNNNNNNNNNNNNN  NNNNNNNNNNNNNNNNNNNN    1
S    1782 hCoV-19/South Africa/KRISP-K004306/2020|EPI_ISL_66    ····················  ····················  ····················  ····················  ·NNNNNNNNNNNNNNNNNNN  NNNNNNNNNNNNNNNNNNNN    1
S    1815 hCoV-19/Spain/MD-IBV-99008628/2020|EPI_ISL_654202|    ····················  ····················  ····················  ····················  NNNNNNNNNNNNNNNNNNNN  NNNNNNNNNNNNNNNNNNNN    1
S    1819 hCoV-19/Spain/MD-IBV-99008635/2020|EPI_ISL_654206|    ····················  ····················  ····················  ····················  NNNNNNNNNNNNNNNNNNNN  NNNNNNNNNNNNNNNNNNNN    1
S    1821 hCoV-19/Spain/MD-IBV-99008637/2020|EPI_ISL_654208|    ····················  ····················  ····················  ····················  NNNNNNNNNNNNNNNNNNNN  ·············NN·····    1
S    1822 hCoV-19/Spain/MD-IBV-99008638/2020|EPI_ISL_654209|    ····················  ····················  ····················  ····················  NNNNNNN·············  ····················    1
S    1823 hCoV-19/Spain/MD-IBV-99008642/2020|EPI_ISL_654210|    ····················  ····················  ····················  ····················  NNNNNNNNNNNNNNNNNNNN  ····················    1
S    1824 hCoV-19/Spain/MD-IBV-99008643/2020|EPI_ISL_654211|    ····················  ····················  ····················  ····················  NNNNNNNNNNNNNNNNNNNN  NNNNNNNNNNNNNNNNNNNN    1
S    1825 hCoV-19/Spain/MD-IBV-99008644/2020|EPI_ISL_654212|    ····················  ····················  ····················  ····················  NNNNNNNNNNNNNNNNNNNN  NNNNNNNNNNNNNNNNNNNN    1
S    1826 hCoV-19/Spain/MD-IBV-99008647/2020|EPI_ISL_654213|    ····················  ····················  ····················  ····················  NNNNNNNNNNNNNNNNNNNN  NNNNNNNNNNNNNNNNNNNN    1
S    1827 hCoV-19/Spain/MD-IBV-99008648/2020|EPI_ISL_654214|    ····················  ····················  ····················  ····················  NNNNNNNNNNNNNNNNNNNN  NNNNNNNNNNNNNNNNNNNN    1
S    1833 hCoV-19/Spain/MD-IBV-99008662/2020|EPI_ISL_654223|    ····················  ····················  ····················  ····················  NNNNNNNNNNNNNNNNNNNN  NNNNNNNNNNNNNNNNNNNN    1
S    1836 hCoV-19/Spain/MD-IBV-99008667/2020|EPI_ISL_654228|    ····················  ····················  ····················  ····················  NNNNNNNNNNNNNNNNNNNN  NNNNNNNNNNNNNNNNNNNN    1
S    1837 hCoV-19/Spain/MD-IBV-99008668/2020|EPI_ISL_654229|    ····················  ····················  ····················  ····················  NNNNNNNNNNNNNNNNNNNN  NNNNNNNNNNNNNNNNNNNN    1
S    1840 hCoV-19/Spain/MD-IBV-99008677/2020|EPI_ISL_654232|    ····················  ····················  ····················  ····················  NNNNNNNNNNNNNNNNNNNN  NNNNNNNNNNNNNNNNNNNN    1
S    1841 hCoV-19/Spain/MD-IBV-99008678/2020|EPI_ISL_654233|    ····················  ····················  ····················  ····················  NNNNNNNNNNNNNNNNNNNN  NNNNNNNNNNNNNNNNNNNN    1
S    1843 hCoV-19/Spain/MD-IBV-99008680/2020|EPI_ISL_654235|    ····················  ····················  ····················  ····················  NNNNNNNNNNNNNNNNNNNN  NNNNNNNNNNNNNNNNNNNN    1
S    1844 hCoV-19/Spain/MD-IBV-99008681/2020|EPI_ISL_654236|    ····················  ····················  ····················  ····················  NNNNNNNNNNNNNNNNNNNN  NNNNNNNNNNNNNNNNNNNN    1
S    1845 hCoV-19/Spain/MD-IBV-99008682/2020|EPI_ISL_654237|    ····················  ····················  ····················  ····················  NNNNNNNNNNNNNNNNNNNN  NNNNNNNNNNNNNNNNNNNN    1
S    1846 hCoV-19/Spain/MD-IBV-99008683/2020|EPI_ISL_654238|    ····················  ····················  ····················  ····················  NNNNNNNNNNNNNNNNNNNN  NNNNNNNNNNNNNNNNNNNN    1
S    1847 hCoV-19/Spain/MD-IBV-99008684/2020|EPI_ISL_654239|    ····················  ····················  ····················  ····················  ····················  NNNNNNNNNNNNNNN····N    1
S    1850 hCoV-19/Spain/MD-IBV-99008688/2020|EPI_ISL_654242|    ····················  ····················  ····················  ····················  NNNNNNNNNNNNNNNNNNNN  NNNNNNNNNNNNNNNNNNNN    1
S    1851 hCoV-19/Spain/MD-IBV-99008689/2020|EPI_ISL_654243|    ····················  ····················  ····················  ····················  NNNNNNNNNNNNNNNNNNNN  NNNNNNNNNNNNNNNNNNNN    1
S    1852 hCoV-19/Spain/MD-IBV-99008690/2020|EPI_ISL_654244|    ····················  ····················  ····················  ····················  NNNNNNNNNNNNNNNNNNNN  NNNNNNNNNNNNNNNNNNNN    1
S    1853 hCoV-19/Spain/MD-IBV-99008691/2020|EPI_ISL_654245|    ····················  ····················  ····················  ····················  NNNNNNNNNNNNNNNNNNNN  NNNNNNNNNNNNNNNNNNNN    1
S    1854 hCoV-19/Spain/MD-IBV-99008692/2020|EPI_ISL_654246|    ····················  ····················  ····················  ····················  NNNNNNNNNNNNNNNNNNNN  NNNNNNNNNNNNNNNNNNNN    1
S    1855 hCoV-19/Spain/MD-IBV-99008694/2020|EPI_ISL_654247|    ····················  ····················  ····················  ····················  NNNNNNNNNNNN········  ····················    1
S    1856 hCoV-19/Spain/MD-IBV-99008721/2020|EPI_ISL_654277|    ····················  ····················  ····················  ····················  NNNNNNNNNNNNNNNNNNNN  NNNNNNNNNNNNNNNNNNNN    1
S    1857 hCoV-19/Spain/MD-IBV-99008698/2020|EPI_ISL_654278|    ····················  ····················  ····················  ····················  ····················  NNNNNNNNNNNNNNNNNNNN    1
S    1862 hCoV-19/Spain/MD-IBV-99008703/2020|EPI_ISL_654283|    ····················  ····················  ····················  ····················  ·NNNNNNNNNNNNNNNNNNN  NNNNNNNNNNNNNNNNNNNN    1
S    1866 hCoV-19/Spain/MD-IBV-99008709/2020|EPI_ISL_654287|    ····················  ····················  ····················  ····················  NNNNNNNNNNNNNNNNNNNN  NNNNNNNNNNNNNNNNNNNN    1
S    1872 hCoV-19/Spain/MD-IBV-99008716/2020|EPI_ISL_654293|    ····················  ····················  ····················  ····················  ··NNNNN······NNN····  ····················    1
S    1873 hCoV-19/Spain/MD-IBV-99008717/2020|EPI_ISL_654294|    ····················  ····················  ····················  ····················  NNNNNNNNNNNNNNNNNNNN  NNNNNNNNNNNNNNNNNNNN    1
S    1874 hCoV-19/Spain/MD-IBV-99008718/2020|EPI_ISL_654295|    ····················  ····················  ····················  ····················  NNNNNNNNNNNNNNNNNNNN  NNNNNNNNNNNNNNNNNNNN    1
S    1876 hCoV-19/Spain/MD-IBV-99008722/2020|EPI_ISL_654297|    ····················  ····················  ····················  ····················  NNNNNNNNNNNNNNNNNNNN  NNNNNNNNNNNNNNNNNNNN    1
S    1878 hCoV-19/Spain/MD-IBV-99008725/2020|EPI_ISL_654299|    ····················  ····················  ····················  ····················  NNNNNNNNNNNNNNNNNNNN  NNNNNNNNNNNNNNNNNNNN    1
S    2049 hCoV-19/England/NORW-F7EA8/2020|EPI_ISL_651100|202    ····················  ·A··················  ····················  ····················  NNNNNNNNNNNNNNNNNNNN  NNNNNNNNNNNNNNNNNNNN    1
S    2050 hCoV-19/England/NORW-F7EF3/2020|EPI_ISL_651101|202    ····················  ·A··················  ····················  ····················  NNNNNNNNNNNNNNNNNNNN  NNNNNNNNNNNNNNNNNNNN    1
S    2051 hCoV-19/England/NORW-F7F2D/2020|EPI_ISL_651102|202    ····················  ·A··················  ····················  ····················  ····················  NNNNNNNNNNNNNNNNNNNN    1
S    2133 hCoV-19/Colombia/DC-INS-2624/2020|EPI_ISL_653746|2    ····················  ····················  ····················  ····················  NNNNNNNNNNNNNNNNNNNN  NNNNNNNNNNNNNNNNNNNN    1
S    2134 hCoV-19/Colombia/DC-INS-2640/2020|EPI_ISL_653747|2    ····················  ····················  ····················  ····················  NNNNNNNNNNNNNNNNNNNN  NNNNNNNNNNNNNNNNNNNN    1
S    2139 hCoV-19/Colombia/DC-INS-3569/2020|EPI_ISL_653755|2    ····················  ····················  ····················  ····················  NNNNNNNNNNNNNNNNNNNN  NNNNNNNNNNNNNNNNNNNN    1
S    2141 hCoV-19/Colombia/ATL-INS-06/2020|EPI_ISL_653757|20    ····················  ··········A·········  ····················  ····················  NNNNNNNNNNNNNNNNNNNN  NNNNNNNNNNNNNNNNNNNN    1
S    2322 hCoV-19/England/NORW-F1ECC/2020|EPI_ISL_652348|202    ····················  ····················  ····················  ····················  NNNNNNNNNNNNNNNNNNNN  NNNNNNNNNNNNNNNNNNNN    1
S    2326 hCoV-19/England/NORW-F693F/2020|EPI_ISL_652352|202    ····················  ····················  ····················  ····················  NNNNNNNNNNNN········  ····················    1
S    2328 hCoV-19/England/NORW-F6A96/2020|EPI_ISL_652354|202    ····················  ····················  ····················  ····················  ····················  NNNNN···············    1
S    2330 hCoV-19/England/NORW-F6B1B/2020|EPI_ISL_652356|202    ····················  ····················  ····················  ····················  ····················  NNNNNNNNNNNNNNN·····    1
S    2333 hCoV-19/England/NORW-F6DAC/2020|EPI_ISL_652359|202    ····················  ····················  ····················  ····················  NNNN················  NNNNNNNNNNNNNNNNN·NN    1
S    2334 hCoV-19/England/NORW-F79D4/2020|EPI_ISL_652360|202    ····················  ····················  ····················  ····················  NNNNNNNNNNNNNNNNNNNN  NNNNNNNNNNNNNNNNNNNN    1
S    2335 hCoV-19/England/NORW-F7A0E/2020|EPI_ISL_652361|202    ····················  ····················  ····················  ····················  NNNNNNNNNNNNNNNNNNNN  NNN·················    1
S    2355 hCoV-19/England/NOTT-11BAF4/2020|EPI_ISL_652381|20    ····················  ····················  ····················  ····················  NNNNNNNNNNNNNNNNNNNN  NNNNNNNNNNNNNNNNNNNN    1
S    2356 hCoV-19/England/NOTT-11BBC4/2020|EPI_ISL_652382|20    ····················  ····················  ····················  ····················  NNNNNNNNNNNNNNNNNNNN  NNNNNNNNNNNNNNNNNNNN    1
S    2365 hCoV-19/England/NOTT-11BECB/2020|EPI_ISL_652391|20    ····················  ····················  ····················  ····················  NNNNNNNNNNNNNNNNNNNN  NNNNNNNNNNNNNNNNNNNN    1
S    2564 hCoV-19/England/NORW-F7ED5/2020|EPI_ISL_651093|202    ····················  ····················  ····················  ····················  NNNNNNNNNNNNNNNNNNNN  NNNNNNNNNNNNNNNNNNNN    1
S    2856 hCoV-19/England/MILK-B39A6F/2020|EPI_ISL_652302|20    ····················  ····················  ····················  ····················  NNNNNNNNNNNNNNNNNNNN  NNNNNNNNNNNNNNNNNNNN    1
S    2857 hCoV-19/England/MILK-B39B02/2020|EPI_ISL_652303|20    ····················  ····················  ····················  ····················  NNNNNNNNNNNNNNNNNNNN  NNNNNNNNNNNNNNNNNNNN    1
S    3287 hCoV-19/France/OCC-SC515/2020|EPI_ISL_660704|2020-    ····················  ······NNNNN·········  ····················  ····················  ····················  ····················    1
S    4085 hCoV-19/South Africa/KRISP-EC-K004564/2020|EPI_ISL    ····················  ····················  ····················  ····················  ····················  NNNNNNNNNNNNNNNNNNNN    1
S    4086 hCoV-19/South Africa/KRISP-EC-K004565/2020|EPI_ISL    ····················  ····················  ····················  ····················  NNNNNNNNNNNNNNNNNNNN  NNNNNNNNNNNNNNNNNNNN    1
S    4087 hCoV-19/South Africa/KRISP-EC-K004572/2020|EPI_ISL    ····················  ····················  ····················  ····················  ··NNNNNNNNNNNNNNNNNN  NNNNNNNNNNNNNNNNNNNN    1
S    4109 hCoV-19/Scotland/EDB8870/2020|EPI_ISL_652012|2020-    ····················  ····················  ····················  ····················  NNNNNNNNNNNNNNNNNNNN  NNNNNNNNNNNNNNNNNNNN    1
S    4117 hCoV-19/Scotland/EDB8913/2020|EPI_ISL_652020|2020-    ····················  ····················  ····················  ····················  NNNNNNNNNNNNNNNNNNNN  NNNNNNNNNNNNNNNNNNNN    1
S    4118 hCoV-19/Scotland/EDB8918/2020|EPI_ISL_652021|2020-    ····················  ····················  ····················  ····················  NNNNNNNNNNNNNNNNNNNN  NNNNNNNNNNNNNNNNNNNN    1
S    4122 hCoV-19/Scotland/EDB8968/2020|EPI_ISL_652025|2020-    ····················  ····················  ····················  ····················  NNNNNNNNNNNNNNNNNNNN  NNNNNNNNNNNNNNNNNNNN    1
S    4123 hCoV-19/Scotland/EDB8970/2020|EPI_ISL_652026|2020-    ····················  ····················  ····················  ····················  NNNNNNNNNNNNNNNNNNNN  NNNNNNNNNNNNNNNNNNNN    1
```

| | | | | | | | | | |
|---|---|---|---|---|---|---|---|---|---|
| S | 4124 | hCoV-19/Scotland/EDB8972/2020\|EPI_ISL_652027\|2020- | ···················· | ···················· | ···················· | ···················· | NNNNNNNNNNNNNNNNNNNN | NNNNNNNNNNNNNNNNNNNN | 1 |
| S | 4125 | hCoV-19/Scotland/EDB8987/2020\|EPI_ISL_652028\|2020- | ···················· | ···················· | ···················· | ···················· | NNNNNNNNNNNNNNNNNNNN | NNNNNNNNNNNNNNNNNNNN | 1 |
| S | 4126 | hCoV-19/Scotland/EDB8990/2020\|EPI_ISL_652029\|2020- | ···················· | ···················· | ···················· | ···················· | NNNNNNNNNNNNNNNNNNNN | NNNNNNNNNNNNNNNNNNNN | 1 |
| S | 4130 | hCoV-19/Scotland/EDB9014/2020\|EPI_ISL_652033\|2020- | ···················· | ···················· | ···················· | ···················· | NNNNNNNNNNNNNNNNNNNN | NNNNNNNNNNNNNNNNNNNN | 1 |
| S | 4131 | hCoV-19/Scotland/EDB9018/2020\|EPI_ISL_652034\|2020- | ···················· | ···················· | ···················· | ···················· | NNNNNNNNNNNNNNNNNNNN | NNNNNNNNNNNNNNNNNNNN | 1 |
| S | 4135 | hCoV-19/Scotland/EDB9048/2020\|EPI_ISL_652038\|2020- | ···················· | ···················· | ···················· | ···················· | NNNNNNNNNNNNNNNNNNNN | NNNNNNNNNNNNNNNNNNNN | 1 |
| S | 4136 | hCoV-19/Scotland/EDB9050/2020\|EPI_ISL_652039\|2020- | ···················· | ···················· | ···················· | ···················· | NNNNNNNNNNNNNNNNNNNN | NNNNNNNNNNNNNNNNNNNN | 1 |
| S | 4137 | hCoV-19/Scotland/EDB9051/2020\|EPI_ISL_652040\|2020- | ···················· | ···················· | ···················· | ···················· | NNNNNNNNNNNNNNNNNNNN | NNNNNNNNNNNNNNNNNNNN | 1 |
| S | 4148 | hCoV-19/Scotland/EDB9231/2020\|EPI_ISL_652051\|2020- | ···················· | ···················· | ···················· | ···················· | NNNNNNNNNNNNNNNNNNNN | NNNNNNNNNNNNNNNNNNNN | 1 |
| S | 4155 | hCoV-19/Scotland/EDB9456/2020\|EPI_ISL_652058\|2020- | ···················· | ···················· | ···················· | ···················· | NNNNNNNNNNNNNNNNNNNN | NNNNNNNNNNNNNNNNNNNN | 1 |
| S | 4168 | hCoV-19/USA/FL-BPHL-2180/2020\|EPI_ISL_653333\|2020- | ···················· | ···················· | ···················· | ···················· | NNNNNNNNNNNNNNNNNNNN | NNNNNNNNNNNNNNNNNNNN | 1 |
| S | 4217 | hCoV-19/Spain/AS-IBV-98006586/2020\|EPI_ISL_654635\| | ···················· | ···················· | ···················· | ···················· | NNNNNNNNNNNNNNNNNNNN | NNNNNNNNNNNNNNNNNNNN | 1 |
| S | 4218 | hCoV-19/Spain/AS-IBV-98006587/2020\|EPI_ISL_654636\| | ···················· | ···················· | ···················· | ···················· | NNNNNNNNNNNNNNNNNNNN | NNNNNNNNNNNNNNNNNNNN | 1 |
| S | 4416 | hCoV-19/England/BIRM-66E59/2020\|EPI_ISL_651597\|202 | ···················· | ·A·················· | ···················· | ···················· | NNNNNNNNNNNNNNNNNNNN | NNNNNNNNNNNNNNNNNNNN | 1 |
| S | 4417 | hCoV-19/England/BIRM-66E68/2020\|EPI_ISL_651598\|202 | ···················· | ···················· | ···················· | ···················· | NNNNNNNNNNNNNNNNNNNN | NNNNNNNNNNNNNNNNNNNN | 1 |
| S | 4418 | hCoV-19/England/BIRM-66E77/2020\|EPI_ISL_651599\|202 | ···················· | ···················· | ···················· | ···················· | NNNNNNNNNNNNNNNNNNNN | NNNNNNNNNNNNNNNNNNNN | 1 |
| S | 4609 | hCoV-19/Equatorial Guinea/44402/2020\|EPI_ISL_64917 | ···················· | ···················· | ···················· | ···················· | NNNNNNNNNNNNNNNNNNNN | NNNNNNNNNNNNNNNNNNNN | 1 |
| S | 4610 | hCoV-19/Equatorial Guinea/44989/2020\|EPI_ISL_64917 | ···················· | ···················· | ···················· | ···················· | NNNNNNNNNNNNNNNNNNNN | NNNNNNNNNNNNNNNNNNNN | 1 |
| S | 4611 | hCoV-19/Equatorial Guinea/46215/2020\|EPI_ISL_64917 | ···················· | ···················· | ···················· | ···················· | NNNNNNNNNNNNNNNNNNNN | NNNNNNNNNNNNNNNNNNNN | 1 |
| S | 4668 | hCoV-19/Scotland/EDB9056/2020\|EPI_ISL_651523\|2020- | ···················· | ···················· | ···················· | ···················· | NNNNNNNNNNNNNNNNNNNN | NNNNNNNNNNNNNNNNNNNN | 1 |
| S | 4669 | hCoV-19/Scotland/EDB9175/2020\|EPI_ISL_651524\|2020- | ···················· | ···················· | ···················· | ···················· | NNNNNNNNNNNNNNNNNNNN | NNNNNNNNNNNNNNNNNNNN | 1 |
| S | 4821 | hCoV-19/Mexico/GRO-InDRE-93/2020\|EPI_ISL_658901\|20 | ···················· | ···················· | ···················· | ···················· | NNNNNNNNNNNNNNNNNNNN | NNNNNNNNNNNNNNNNNNNN | 1 |
| S | 4866 | hCoV-19/Equatorial Guinea/12779/2020\|EPI_ISL_64915 | ···················· | ···················· | ···················· | ···················· | NNNNNNNNNNNNNNNNNNNN | NNNNNNNNNNNNNNNNNNNN | 1 |
| S | 4868 | hCoV-19/Equatorial Guinea/27213/2020\|EPI_ISL_64916 | ···················· | ···················· | ···················· | ···················· | NNNNNNN············· | ···················· | 1 |
| S | 5126 | hCoV-19/USA/MI-UM-10036634235/2020\|EPI_ISL_650036\| | ···················· | ·A·················· | ···················· | ···················· | NNNNNNNNNNNNNNNNNNNN | NNNNNNNNNNNNNNNNNNNN | 1 |
| S | 5127 | hCoV-19/USA/MI-UM-10036632926/2020\|EPI_ISL_650037\| | ···················· | ···················· | ···················· | ···················· | ··NNNNNNNNNNNNNNNNNN | ···················· | 1 |
| S | 5128 | hCoV-19/USA/MI-UM-10036631770/2020\|EPI_ISL_650038\| | ···················· | ···················· | ···················· | ···················· | NNNNNNN············· | ···················· | 1 |
| S | 5130 | hCoV-19/USA/MI-UM-10036633059/2020\|EPI_ISL_650041\| | ···················· | ···················· | ···················· | ···················· | NNNNNNNNNNNNNNNNNNNN | NNNNNNNNNNNNNNNNNNNN | 1 |
| S | 5134 | hCoV-19/USA/MI-UM-10036633901/2020\|EPI_ISL_650046\| | ···················· | ·····A·············· | ···················· | ···················· | NNNNNNNNNNNNNNNNNNNN | ···················· | 1 |
| S | 5135 | hCoV-19/USA/MI-UM-10036672506/2020\|EPI_ISL_650047\| | ···················· | ···················· | ···················· | ···················· | NNNNNNNNNNNNNNNNNNNN | ···················· | 1 |
| S | 5138 | hCoV-19/USA/MI-UM-10036672489/2020\|EPI_ISL_650050\| | ···················· | ···················· | ···················· | ···················· | NNNNNNNNNNNNNNNNNNNN | ···················· | 1 |
| S | 5140 | hCoV-19/USA/MI-UM-10036649695/2020\|EPI_ISL_650052\| | ···················· | ···················· | ···················· | ···················· | NNNNNNN············· | ···················· | 1 |
| S | 5151 | hCoV-19/USA/MI-UM-10036652302/2020\|EPI_ISL_650065\| | ···················· | ···················· | ···················· | ···················· | NNNNNNNNNNNNNNNNNNNN | ···················· | 1 |
| S | 5153 | hCoV-19/USA/MI-UM-10036648955/2020\|EPI_ISL_650067\| | ···················· | ···················· | ···················· | ···················· | NNNNNNNNNNNNNNNNNNNN | NNNNNNNNNNNNNNNNNNNN | 1 |
| S | 5154 | hCoV-19/USA/MI-UM-10036660941/2020\|EPI_ISL_650068\| | ···················· | ···················· | ···················· | ···················· | NNNNNNNNNNNNNNNNNNNN | NNNNNNNNNNNNNNNNNNNN | 1 |
| S | 5156 | hCoV-19/USA/MI-UM-10036648964/2020\|EPI_ISL_650071\| | ···················· | ···················· | ···················· | ···················· | NNNNNNNNNNNNNNNNNNNN | NNNNNNNNNNNNNNNNNNNN | 1 |
| S | 5157 | hCoV-19/USA/MI-UM-10036674506/2020\|EPI_ISL_650073\| | ···················· | ···················· | ···················· | ···················· | NNNNNNNNNNNNNNNNNNNN | ···················· | 1 |
| S | 5158 | hCoV-19/USA/MI-UM-10036673508/2020\|EPI_ISL_650075\| | ···················· | ···················· | ···················· | ···················· | NNNNNNNNNNNNNNNNNNNN | ···················· | 1 |
| S | 5159 | hCoV-19/USA/MI-UM-10036644998/2020\|EPI_ISL_650076\| | ···················· | ···················· | ···················· | ···················· | NNNNNNNNNNNNNNNNNNNN | NNNNNNNNNNNNNNNNNNNN | 1 |
| S | 5161 | hCoV-19/USA/MI-UM-10036648850/2020\|EPI_ISL_650078\| | ···················· | ···················· | ···················· | ···················· | NNNNNNNNNNNNNNNNNNNN | NNNNNNNNNNNNNNNNNNNN | 1 |
| S | 5162 | hCoV-19/USA/MI-UM-10036662641/2020\|EPI_ISL_650079\| | ···················· | ···················· | ···················· | ···················· | NNNNNNNNNNNNNNNNNNNN | NNNNNNNNNNNNNNNNNNNN | 1 |
| S | 5163 | hCoV-19/USA/MI-UM-10036660976/2020\|EPI_ISL_650083\| | ···················· | ···················· | ···················· | ···················· | NNNNNNNNNNNNNNNNNNNN | NNNNNNNNNNNNNNNNNNNN | 1 |
| S | 5164 | hCoV-19/USA/MI-UM-10036682610/2020\|EPI_ISL_650085\| | ···················· | ···················· | ···················· | ···················· | NNNNNNNNNNNNNNNNNNNN | NNNNNNNNNNNNNNNNNNNN | 1 |
| S | 5165 | hCoV-19/USA/MI-UM-10036693063/2020\|EPI_ISL_650087\| | ···················· | ···················· | ···················· | ···················· | NNNNNNNNNNNNNNNNNNNN | NNNNNNNNNNNNNNNNNNNN | 1 |
| S | 5166 | hCoV-19/USA/MI-UM-10036687972/2020\|EPI_ISL_650088\| | ···················· | ···················· | ···················· | ···················· | NNNNNNNNNNNNNNNNNNNN | ···················· | 1 |
| S | 5167 | hCoV-19/USA/MI-UM-10036673492/2020\|EPI_ISL_650089\| | ···················· | ············A······· | ···················· | ···················· | NNNNNNNNNNNNNNNNNNNN | NNNNNNNNNNNNNNNNNNNN | 1 |
| S | 5168 | hCoV-19/USA/MI-UM-10036687345/2020\|EPI_ISL_650090\| | ···················· | ············A······· | ···················· | ···················· | NNNNNNNNNNNNNNNNNNNN | ···················· | 1 |
| S | 5169 | hCoV-19/USA/MI-UM-10036667418/2020\|EPI_ISL_650091\| | ···················· | ···················· | ···················· | ···················· | NNNNNNNNNNNNNNNNNNNN | NNNNNNNNNNNNNNNNNNNN | 1 |
| S | 5170 | hCoV-19/USA/MI-UM-10036671254/2020\|EPI_ISL_650092\| | ···················· | ············A······· | ···················· | ···················· | ············NNNNNNNN | ···················· | 1 |
| S | 5172 | hCoV-19/USA/MI-UM-10036696400/2020\|EPI_ISL_650094\| | ···················· | ···················· | ···················· | ···················· | ······NNNNNNNNNNNNNN | ···················· | 1 |
| S | 5173 | hCoV-19/USA/MI-UM-10036688546/2020\|EPI_ISL_650095\| | ···················· | ···················· | ···················· | ···················· | NNNNNNNNNNNNNNNNNNNN | ···················· | 1 |
| S | 5174 | hCoV-19/USA/MI-UM-10036661877/2020\|EPI_ISL_650096\| | ···················· | ···················· | ···················· | ···················· | NNNNNNNNNNNNNNNNNNNN | NNNNNNNNNNNNNNNNNNNN | 1 |
| S | 5176 | hCoV-19/USA/MI-UM-10036669656/2020\|EPI_ISL_650098\| | ···················· | ···················· | ···················· | ···················· | NNNNNNNNNNNNNNNNNNNN | ···················· | 1 |
| S | 5178 | hCoV-19/USA/MI-UM-10036669328/2020\|EPI_ISL_650100\| | ···················· | ·A·················· | ···················· | ···················· | NNNNNNNNNNNNNNNNNNNN | NNNNNNNNNNNNNNNNNNNN | 1 |
| S | 5180 | hCoV-19/USA/MI-UM-10036671867/2020\|EPI_ISL_650102\| | ···················· | ···················· | ···················· | ···················· | NNNNNNNNNNNNNNNNNNNN | NNNNNNNNNNNNNNNNNNNN | 1 |
| S | 5181 | hCoV-19/USA/MI-UM-10036691262/2020\|EPI_ISL_650104\| | ···················· | ···················· | ···················· | ···················· | NNNNNNNNNNNNNNNNNNNN | NNNNNNNNNNNNNNNNNNNN | 1 |
| S | 5182 | hCoV-19/USA/MI-UM-10036683197/2020\|EPI_ISL_650106\| | ···················· | ·A·················· | ···················· | ···················· | NNNNNNNNNNNNNNNNNNNN | NNNNNNNNNNNNNNNNNNNN | 1 |
| S | 5183 | hCoV-19/England/BIRM-66E86/2020\|EPI_ISL_651413\|202 | ···················· | ···················· | ···················· | ···················· | NNNNNNNNNNNNNNNNNNNN | NNNNNNNNNNNNNNNNNNNN | 1 |
| S | 5378 | hCoV-19/USA/MI-UM-10036589901/2020\|EPI_ISL_650002\| | ···················· | ···················· | ···················· | ···················· | NNNNNNNNNNNNNNNNNNNN | ···················· | 1 |
| S | 5379 | hCoV-19/USA/MI-UM-10036606626/2020\|EPI_ISL_650016\| | ···················· | ···················· | ···················· | ···················· | NNNNNNNNNNNNNNNNNNNN | NNNNNNNNNNNNNNNNNNNN | 1 |
| S | 5380 | hCoV-19/USA/MI-UM-10036608175/2020\|EPI_ISL_650017\| | ···················· | ···················· | ···················· | ···················· | NNNNNNNNNNNNNNNNNNNN | NNNNNNNNNNNNNNNNNNNN | 1 |
| S | 5381 | hCoV-19/USA/MI-UM-10036608612/2020\|EPI_ISL_650018\| | ···················· | ···················· | ···················· | ···················· | NNNNNNNNNNNNNNNNNNNN | NNNNNNNNNNNNNNNNNNNN | 1 |
| S | 5383 | hCoV-19/USA/MI-UM-10036630690/2020\|EPI_ISL_650020\| | ···················· | ·····A·············· | ···················· | ···················· | ·NNNNNNNNNNNNNNNNNNN | ···················· | 1 |
| S | 5384 | hCoV-19/USA/MI-UM-10036619785/2020\|EPI_ISL_650021\| | ···················· | ···················· | ···················· | ···················· | NNNNNNNNNNNNNNNNNNNN | ···················· | 1 |
| S | 5386 | hCoV-19/USA/MI-UM-10036626781/2020\|EPI_ISL_650024\| | ···················· | ···················· | ···················· | ···················· | NNNNNNNNNNNNNNNNNNNN | ···················· | 1 |
| S | 5390 | hCoV-19/USA/MI-UM-10036652539/2020\|EPI_ISL_650034\| | ···················· | ···················· | ···················· | ···················· | NNNNNNNNNNNNNNNNNNNN | ···················· | 1 |
| S | 5391 | hCoV-19/USA/MI-UM-10036619926/2020\|EPI_ISL_650035\| | ···················· | ···················· | ···················· | ···················· | NNNNNNNNNNNNNNNNNNNN | ···················· | 1 |
| S | 5633 | hCoV-19/USA/MI-UM-10036614249/2020\|EPI_ISL_650003\| | ···················· | ···················· | ···················· | ···················· | NNNNNNNNNNNNNNNNNNNN | NNNNNNNNNNNNNNNNNNNN | 1 |
| S | 5634 | hCoV-19/USA/MI-UM-10036605246/2020\|EPI_ISL_650004\| | ···················· | ···················· | ···················· | ···················· | NNNNNNNNNNNNNNNNNNNN | NNNNNNNNNNNNNNNNNNNN | 1 |
| S | 5635 | hCoV-19/USA/MI-UM-10036613887/2020\|EPI_ISL_650006\| | ···················· | ···················· | ···················· | ···················· | NNNNNNNNNNNNNNNNNNNN | ···················· | 1 |
| S | 5636 | hCoV-19/USA/MI-UM-10036587365/2020\|EPI_ISL_650007\| | ···················· | ···················· | ···················· | ···················· | NNNNNNNNNNNNNNNNNNNN | NNNNNNNNNNNNNNNNNNNN | 1 |
| S | 5639 | hCoV-19/USA/MI-UM-10036592138/2020\|EPI_ISL_650012\| | ···················· | ···················· | ···················· | ···················· | NNNNNNNNNNNNNNNNNNNN | NNNNNNNNNNNNNNNNNNNN | 1 |
| S | 5738 | hCoV-19/England/204441477/2020\|EPI_ISL_653972\|2020 | ···················· | ···················· | ···················· | NNNNNNNNNNNNNNNNNNNN | ···················· | ···················· | 1 |
| S | 5891 | hCoV-19/Wales/PHWC-48A1E9/2020\|EPI_ISL_652535\|2020 | ···················· | ···················· | ···················· | ···················· | NNN················· | NNNNNNNNNNNNNNN····· | 1 |
| S | 5922 | hCoV-19/Wales/PHWC-48A556/2020\|EPI_ISL_652568\|2020 | ···················· | ···················· | ···················· | ···················· | ···NNNNNNNNNNNNNNNNN | ···················· | 1 |
| S | 5924 | hCoV-19/Wales/PHWC-48A592/2020\|EPI_ISL_652570\|2020 | ···················· | ···················· | ···················· | ···················· | NNNNNNNNNNNNNNNNNNNN | NNNNNNNNNNNNNNNNNNNN | 1 |
| S | 6189 | hCoV-19/Wales/PHWC-489E7A/2020\|EPI_ISL_652500\|2020 | ···················· | ···················· | ···················· | ···················· | NNNNNNNNNNNNNNNNNNNN | ···················· | 1 |
| S | 6401 | hCoV-19/Scotland/QEUH-AE20A0/2020\|EPI_ISL_649650\|2 | ···················· | ···················· | ···················· | NNNNNNNNNNNNNNNNNNNN | ···················· | ···················· | 1 |
| S | 6431 | hCoV-19/Scotland/EDB8569/2020\|EPI_ISL_651962\|2020- | ···················· | ···················· | ···················· | ···················· | NNNNNNNNNNNNNNNNNNNN | NNNNNNNNNNNNNNNNNNNN | 1 |
| S | 6432 | hCoV-19/Scotland/EDB8572/2020\|EPI_ISL_651963\|2020- | ···················· | ···················· | ···················· | ···················· | NNNNNNNNNNNNNNNNNNNN | NNNNNNNNNNNNNNNNNNNN | 1 |

```
S    6433 hCoV-19/Scotland/EDB8583/2020|EPI_ISL_651964|2020-   ····················  ····················  ····················  ····················  NNNNNNNNNNNNNNNNNNNN  NNNNNNNNNNNNNNNNNNNN    1
S    6434 hCoV-19/Scotland/EDB8584/2020|EPI_ISL_651965|2020-   ····················  ····················  ····················  ····················  NNNNNNNNNNNNNNNNNNNN  NNNNNNNNNNNNNNNNNNNN    1
S    6437 hCoV-19/Scotland/EDB8587/2020|EPI_ISL_651968|2020-   ····················  ····················  ····················  ····················  NNNNNNNNNNNNNNNNNNNN  NNNNNNNNNNNNNNNNNNNN    1
S    6439 hCoV-19/Scotland/EDB8590/2020|EPI_ISL_651970|2020-   ····················  ····················  ····················  ····················  NNNNNNNNNNNNNNNNNNNN  NNNNNNNNNNNNNNNNNNNN    1
S    6440 hCoV-19/Scotland/EDB8591/2020|EPI_ISL_651971|2020-   ····················  ····················  ····················  ····················  NNNNNNNNNNNNNNNNNNNN  NNNNNNNNNNNNNNNNNNNN    1
S    6441 hCoV-19/Scotland/EDB8597/2020|EPI_ISL_651974|2020-   ····················  ····················  ····················  ····················  NNNNNNNNNNNNNNNNNNNN  NNNNNNNNNNNNNNNNNNNN    1
S    6457 hCoV-19/Scotland/EDB8774/2020|EPI_ISL_651994|2020-   ····················  ····················  ····················  ····················  NNNNNNNNNNNNNNNNNNNN  NNNNNNNNNNNNNNNNNNNN    1
S    6759 hCoV-19/Spain/MD-IBV-99008560/2020|EPI_ISL_654100|   ····················  ····················  ····················  ····················  NNNNNNNNNNNNNNNNNNNN  NNNNNNNNNNNNNNNNNNNN    1
S    6760 hCoV-19/Spain/MD-IBV-99008561/2020|EPI_ISL_654101|   ····················  ····················  ····················  ····················  NNNNNNN·············  ····················    1
S    6761 hCoV-19/Spain/MD-IBV-99008562/2020|EPI_ISL_654102|   ····················  ····················  ····················  ····················  NNNNNNNNNNNNNNNNNNNN  NNNNNNNNNNNNNNNNNNNN    1
S    6764 hCoV-19/Spain/MD-IBV-99008565/2020|EPI_ISL_654105|   ····················  ····················  ····················  ····················  NNNNNNNNNNNNNNNNNNNN  NNNNNNNNNNNNNNNNNNNN    1
S    6967 hCoV-19/Spain/MD-IBV-99008566/2020|EPI_ISL_654106|   ····················  ····················  ····················  ····················  NNNNNNNNNNNNNNNNNNNN  NNNNNNNNNNNNNNNNNNNN    1
S    6968 hCoV-19/Spain/MD-IBV-99008568/2020|EPI_ISL_654108|   ····················  ····················  ····················  ····················  NNNNNNNNNNNNNNNNNNNN  NN··NNNNNNNNNNN·····    1
S    6969 hCoV-19/Spain/MD-IBV-99008569/2020|EPI_ISL_654109|   ····················  ····················  ····················  ····················  NNNNNNNNNNNNNNNNNNNN  NNNNNNNNNNNNNNNNNNNN    1
S    6971 hCoV-19/Spain/MD-IBV-99008571/2020|EPI_ISL_654111|   ····················  ····················  ····················  ····················  NNNNNNNNNNNNNNNNNNNN  NNNNNNNNNNNNNNN·····    1
S    6973 hCoV-19/Spain/MD-IBV-99008573/2020|EPI_ISL_654113|   ····················  ····················  ····················  ····················  NNNNNNNNNNNNNNNNNNNN  NNNNNNNNNNNNNNNNNNNN    1
S    6975 hCoV-19/Spain/MD-IBV-99008575/2020|EPI_ISL_654115|   ····················  ····················  ····················  ····················  NNNNNNNNNNNNNNNNNNNN  NNNNNNNNNNNNNNN·····    1
S    6976 hCoV-19/Spain/MD-IBV-99008576/2020|EPI_ISL_654116|   ····················  ·····A··············  ····················  ····················  NNNNNNNNNNNNNNNNNNNN  ············NNN····N    1
S    6978 hCoV-19/Spain/MD-IBV-99008578/2020|EPI_ISL_654118|   ····················  ····················  ····················  ····················  NNNNNNNNNNNNNNNNNNNN  NNNNNNNNNNNNNNNNNNNN    1
S    6979 hCoV-19/Spain/MD-IBV-99008579/2020|EPI_ISL_654119|   ····················  ····················  ····················  ····················  NNNNNNNNNNNNNNNNNNNN  NNNNNNNNNNNNNNNNNNNN    1
S    6980 hCoV-19/Spain/MD-IBV-99008580/2020|EPI_ISL_654120|   ····················  ····················  ····················  ····················  NNNNNNNNNNNNNNNNNNNN  NNNNNNNNNNNNNNNNNNNN    1
S    6983 hCoV-19/Spain/MD-IBV-99008583/2020|EPI_ISL_654123|   ····················  ····················  ····················  ····················  NNNNNNNNNNNNNNNNNNNN  NNNNNNNNNNNNNNNNNNNN    1
S    6985 hCoV-19/Spain/MD-IBV-99008586/2020|EPI_ISL_654126|   ····················  ····················  ····················  ····················  NNNNNNNNNNNNNNNNNNNN  NNNNNNNNNNNNNNN·····    1
S    6986 hCoV-19/Spain/MD-IBV-99008587/2020|EPI_ISL_654127|   ····················  ····················  ····················  ····················  NNNNNNNNNNNNNNNNNNNN  NNNNNNNNNNNNNNNNNNNN    1
S    6990 hCoV-19/Spain/MD-IBV-99008591/2020|EPI_ISL_654131|   ····················  ····················  ····················  ····················  NNNNNNNNNNNNNNNNNNNN  NNNNNNNNNNNNNNN·····    1
S    6991 hCoV-19/Spain/MD-IBV-99008592/2020|EPI_ISL_654132|   ····················  ····················  ····················  ····················  NNNNNNNNNNNNNNNNNNNN  NNNNNNNNNNNNNNNNNNNN    1
S    6994 hCoV-19/Spain/MD-IBV-99008596/2020|EPI_ISL_654136|   ····················  ····················  ····················  ····················  NNNNNNNNNNNNNNNNNNNN  NNNNNNNNNNNNNNNNNNNN    1
S    6997 hCoV-19/Spain/MD-IBV-99008600/2020|EPI_ISL_654139|   ····················  ····················  ····················  ····················  NNNNNNNNNNNNNNNNNNNN  NNNNNNNNNNNNNNNNNNNN    1
S    6998 hCoV-19/Spain/MD-IBV-99008790/2020|EPI_ISL_654141|   ····················  ····················  ····················  ····················  NNNNNNNNNNNNNNNNNNNN  NNNNNNNNNNNNNNNNNNNN    1
S    7001 hCoV-19/Spain/MD-IBV-99008793/2020|EPI_ISL_654144|   ····················  ····················  ····················  ····················  NNNNNNN·············  ····················    1
S    7002 hCoV-19/Spain/MD-IBV-99008794/2020|EPI_ISL_654145|   ····················  ····················  ····················  ····················  NNNNNNNNNNNNNNNNNNNN  NNNNNNNNNNNNNNNNNNNN    1
S    7004 hCoV-19/Spain/MD-IBV-99008796/2020|EPI_ISL_654147|   ····················  ····················  ····················  ····················  NNNNNNNNNNNNNNNNNNNN  NNNNNNNNNNNNNNN·····    1
S    7006 hCoV-19/Spain/MD-IBV-99008798/2020|EPI_ISL_654149|   ····················  ····················  ····················  ····················  NNNNNNNNNNNNNNNNNNNN  NNNNNNNNNNNNNNNNNNNN    1
S    7014 hCoV-19/Spain/MD-IBV-99008806/2020|EPI_ISL_654157|   ····················  ····················  ····················  ····················  NNNNNNNNNNNNNNNNNNNN  NNNNNNNNNNNNNNNNNNNN    1
S    7015 hCoV-19/Spain/MD-IBV-99008808/2020|EPI_ISL_654158|   ····················  ····················  ····················  ····················  NNNNNNNNNNNNNNNNNNNN  NNNNNNNNNNNNNNNNNNNN    1
S    7016 hCoV-19/Spain/MD-IBV-99008809/2020|EPI_ISL_654159|   ····················  ····················  ····················  ····················  NNNN················  ····················    1
S    7017 hCoV-19/Spain/MD-IBV-99008811/2020|EPI_ISL_654160|   ····················  ····················  ····················  ····················  ····················  ··········NNNNN·····    1
S    7018 hCoV-19/Spain/MD-IBV-99008812/2020|EPI_ISL_654161|   ····················  ····················  ····················  ····················  NNNNNNNNNNNNNNNNNNNN  NNNNNNNNNNNNNNNNNNNN    1
S    7020 hCoV-19/Spain/MD-IBV-99008601/2020|EPI_ISL_654187|   ····················  ····················  ····················  ····················  NNNNNNNNNNNNNNNNNNNN  NNNNNNNNNNNNNNNNNNNN    1
S    7021 hCoV-19/Spain/MD-IBV-99008602/2020|EPI_ISL_654188|   ····················  ····················  ····················  ····················  NNNNNNNNNNNNNNNNNNNN  ····················    1
S    7022 hCoV-19/Spain/MD-IBV-99008605/2020|EPI_ISL_654189|   ····················  ····················  ····················  ····················  NNNNNNNNNNNNNNNNNNNN  NNNNNNNNNNNNNNNNNNNN    1
S    7023 hCoV-19/Spain/MD-IBV-99008606/2020|EPI_ISL_654190|   ····················  ····················  ····················  ····················  ·NNNNNNNNNNNNNNNNNNN  NNNNNNNNNNNNNNNNNNNN    1
S    7025 hCoV-19/Spain/MD-IBV-99008613/2020|EPI_ISL_654192|   ····················  ····················  ····················  ····················  ·NNNNNN·············  NNNNNNNNNNNNNNN·····    1
S    7029 hCoV-19/Spain/MD-IBV-99008620/2020|EPI_ISL_654196|   ····················  ····················  ····················  ····················  ·NNNNNNNNNNNNNNNNNNN  NNNNNNNNNNNNNNNNNNNN    1
S    7031 hCoV-19/Spain/MD-IBV-99008622/2020|EPI_ISL_654198|   ····················  ····················  ····················  ····················  NNNNNNNNNNNNNNNNNNNN  NNNNNNNNNNNNNNNNNNNN    1
S    7330 hCoV-19/Spain/MD-IBV-99008533/2020|EPI_ISL_654081|   ····················  ····················  ····················  ····················  NNNNNNNNNNNNNNN·····  ····················    1
S    7331 hCoV-19/Spain/MD-IBV-99008534/2020|EPI_ISL_654082|   ····················  ····················  ····················  ····················  NNNNNNNNNNNNNNNNNNNN  NNNNNNNNNNNNNNNNNNNN    1
S    7332 hCoV-19/Spain/MD-IBV-99008535/2020|EPI_ISL_654083|   ····················  ····················  ····················  ····················  NNNNNNNNNNNNNNNNNNNN  NNNNNNNNNNNNNNN·····    1
S    7333 hCoV-19/Spain/MD-IBV-99008536/2020|EPI_ISL_654084|   ····················  ····················  ····················  ····················  NNNNNNNNNNNNNNNNNNNN  NNNNNNNNNNNNNNNNNNNN    1
S    7334 hCoV-19/Spain/MD-IBV-99008539/2020|EPI_ISL_654085|   ····················  ····················  ····················  ····················  NNNNNNNNNNNN········  ····················    1
S    7338 hCoV-19/Spain/MD-IBV-99008543/2020|EPI_ISL_654089|   ····················  ····················  ····················  ····················  NNNNNNNNNNNNNNNNNNNN  ····················    1
S    7339 hCoV-19/Spain/MD-IBV-99008545/2020|EPI_ISL_654090|   ····················  ····················  ····················  ····················  NNNNNNNNNNNNNNNNNNNN  NNNNNNNNNNNNNNNNNNNN    1
S    7340 hCoV-19/Spain/MD-IBV-99008550/2020|EPI_ISL_654092|   ····················  ····················  ····················  ····················  NNNNNNNNNNNN········  ····················    1
S    7341 hCoV-19/Spain/MD-IBV-99008554/2020|EPI_ISL_654095|   ····················  ····················  ····················  ····················  NNNNNNNNNNNN·N······  ····················    1
S    7369 hCoV-19/England/ALDP-B63738/2020|EPI_ISL_656646|20     ····················  NNNNNNNNNNNNNNNNNNNN  ····················  ····················  ····················  ····················    1
S    7436 hCoV-19/Spain/MD-IBV-99008468/2020|EPI_ISL_654054|   ····················  ····················  ····················  ····················  NNNNNNN·············  ····················    1
S    7437 hCoV-19/Spain/MD-IBV-99008469/2020|EPI_ISL_654055|   ····················  ····················  ····················  ····················  NNNNNNNNNNNNNNNNNNNN  NNNNNNNNNNNNNNNNNNNN    1
S    7438 hCoV-19/Spain/MD-IBV-99008472/2020|EPI_ISL_654056|   ····················  ····················  ····················  ····················  NNNNNNNNNNNNNNNNNNNN  NNNNNNNNNNNNNNNNNNNN    1
S    7439 hCoV-19/Spain/MD-IBV-99008473/2020|EPI_ISL_654057|   ····················  ····················  ····················  ····················  NNNNNNNNNNNNNNNNNNNN  NNNNNNNNNNNNNNNNNNNN    1
S    7443 hCoV-19/Spain/MD-IBV-99008479/2020|EPI_ISL_654061|   ····················  ····················  ····················  ····················  NNNNNNNNNNNNNNNNNNNN  ····················    1
S    7445 hCoV-19/Spain/MD-IBV-99008489/2020|EPI_ISL_654063|   ····················  ····················  ····················  ····················  NNNNNNNNNNNNNNNNNNNN  NNNNNNNNNNNNNNNNNNNN    1
S    7446 hCoV-19/Spain/MD-IBV-99008490/2020|EPI_ISL_654064|   ····················  ····················  ····················  ····················  NNNNNNNNNNNNNNNNNNNN  NNNNNNNNNNNNNNNNNNNN    1
S    7447 hCoV-19/Spain/MD-IBV-99008494/2020|EPI_ISL_654065|   ····················  ····················  ····················  ····················  NNNNNNNNNNNNNNNNNNNN  NNNNNNNNNNNNNNNNNNNN    1
S    7448 hCoV-19/Spain/MD-IBV-99008495/2020|EPI_ISL_654066|   ····················  ····················  ····················  ····················  NNNNNNNNNNNNNNNNNNNN  N···················    1
S    7450 hCoV-19/Spain/MD-IBV-99008499/2020|EPI_ISL_654068|   ····················  ····················  ····················  ····················  ·NNNNNNNNNNNNNNNNNNN  NNNNNNNNNNNNNNNNNNNN    1
S    7451 hCoV-19/Spain/MD-IBV-99008500/2020|EPI_ISL_654069|   ····················  ····················  ····················  ····················  NNNNNNNNNNNN········  NNNNN······NNNN·····    1
S    7452 hCoV-19/Spain/MD-IBV-99008503/2020|EPI_ISL_654070|   ····T···············  ····················  ····················  ····················  NNNNNNNNNNNNNNNNNNNN  NNNNNNNNNNNNNNNNNNNN    1
S    7453 hCoV-19/Spain/MD-IBV-99008504/2020|EPI_ISL_654071|   ····················  ····················  ····················  ····················  NNNNNNNNNNNNNNNNNNNN  NNNNNNNNNNNNNNNNNNNN    1
S    7455 hCoV-19/Spain/MD-IBV-99008510/2020|EPI_ISL_654073|   ····················  ····················  ····················  ····················  NNNNNNNNNNNNNNNNNNNN  NNNNNNNNNNNNNNNNNNNN    1
S    7456 hCoV-19/Spain/MD-IBV-99008511/2020|EPI_ISL_654074|   ····················  ····················  ····················  ····················  NNNNNNNNNNNNNNNNNNNN  NNNNNNNNNNNNNNNNN···    1
S    7457 hCoV-19/Spain/MD-IBV-99008517/2020|EPI_ISL_654075|   ····················  ····················  ····················  ····················  NNNNNNNNNNNNNNNNNNNN  NNNNNNNNNNNNNNNNNNNN    1
S    7460 hCoV-19/Spain/MD-IBV-99008524/2020|EPI_ISL_654078|   ····················  ····················  ····················  ····················  NNNNNNNNNNNNNNNNNNNN  NNNNNNNNNNNNNNNNNNNN    1
S    7536 hCoV-19/England/ALDP-B6310D/2020|EPI_ISL_656614|20     ····················  NNNNNNNNNNNNNNNNNNNN  ····················  ····················  ····················  ····················    1
S    7543 hCoV-19/England/ALDP-B63149/2020|EPI_ISL_656621|20     ····················  NNNNNNNNNNNNNNNNNNNN  ····················  ····················  ····················  ····················    1
S    7552 hCoV-19/England/ALDP-B63176/2020|EPI_ISL_656632|20     ····················  NNNNNNNNNNNNNNNNNNNN  ····················  ····················  ····················  ····················    1
S    8035 hCoV-19/England/ALDP-B63765/2020|EPI_ISL_656520|20     ····················  NNNNNNNNNNNNNNNNNNNN  ····················  ····················  ····················  ····················    1
S    8049 hCoV-19/England/ALDP-B61FAB/2020|EPI_ISL_656535|20     ····················  NNNNNNNNNNNNNNNNNNNN  ····················  ····················  ····················  ····················    1
S    8070 hCoV-19/England/ALDP-B637DE/2020|EPI_ISL_656589|20     ····················  NNNNNNNNNNNNNNNNNNNN  ····················  ····················  ····················  ····················    1
S    8232 hCoV-19/England/BIRM-66F29/2020|EPI_ISL_651602|202     ····················  ····················  ····················  ····················  NNNNNNNNNNNNNNNNNNNN  NNNNNNNNNNNNNNNNNNNN    1
S    8233 hCoV-19/England/BIRM-66FA1/2020|EPI_ISL_651603|202     ····················  ····················  ····················  ····················  NNNNNNNNNNNNNNNNNNNN  NNNNNNNNNNNNNNNNNNNN    1
```

| | | | | | | | | | |
|---|---|---|---|---|---|---|---|---|---|
| S | 8234 | hCoV-19/England/BIRM-67016/2020\|EPI_ISL_651604\|202 | ···················· | ···················· | ···················· | ···················· | NNNNNNNNNNNNNNNNNNNN | NNNNNNNNNNNNNNNNNNNN | 1 |
| S | 8237 | hCoV-19/England/BIRM-67131/2020\|EPI_ISL_651607\|202 | ···················· | ···················· | ···················· | ···················· | NNNNNNNNNNNNNNNNNNNN | NNNNNNNNNNNNNNNNNNNN | 1 |
| S | 8239 | hCoV-19/England/BIRM-6715F/2020\|EPI_ISL_651609\|202 | ···················· | ···················· | ···················· | ···················· | NNNNNNNNNNNNNNNNNNNN | NNNNNNNNNNNNNNNNNNNN | 1 |
| S | 8243 | hCoV-19/England/BIRM-674FC/2020\|EPI_ISL_651613\|202 | ···················· | ···················· | ···················· | ···················· | NNNNNNNNNNNNNNNNNNNN | NNNNNNNNNNNNNNNNNNNN | 1 |
| S | 8246 | hCoV-19/England/BIRM-67571/2020\|EPI_ISL_651616\|202 | ···················· | ·A·················· | ···················· | ···················· | NNNNNNNNNNNNNNNNNNNN | NNNNNNNNNNNNNNNNNNNN | 1 |
| S | 8247 | hCoV-19/England/BIRM-67580/2020\|EPI_ISL_651617\|202 | ···················· | ···················· | ···················· | ···················· | NNNNNNNNNNNNNNNNNNNN | NNNNNNNNNNNNNNNNNNNN | 1 |
| S | 8249 | hCoV-19/England/BIRM-675EA/2020\|EPI_ISL_651619\|202 | ···················· | ···················· | ···················· | ···················· | NNNNNNNNNNNNNNNNNNNN | NNNNNNNNNNNNNNNNNNNN | 1 |
| S | 8254 | hCoV-19/England/BIRM-678C3/2020\|EPI_ISL_651624\|202 | ···················· | ···················· | ···················· | ···················· | NNNNNNNNNNNNNNNNNNNN | NNNNNNNNNNNNNNNNNNNN | 1 |
| S | 8257 | hCoV-19/England/BIRM-67A54/2020\|EPI_ISL_651627\|202 | ···················· | ···················· | ···················· | ···················· | NNNNNNNNNNNNNNNNNNNN | NNNNNNNNNNNNNNNNNNNN | 1 |
| S | 8259 | hCoV-19/England/BIRM-67B42/2020\|EPI_ISL_651629\|202 | ···················· | ···················· | ···················· | ···················· | NNNNNNNNNNNNNNNNNNNN | NNNNNNNNNNNNNNNNNNNN | 1 |
| S | 8516 | hCoV-19/England/BIRM-6AB4F/2020\|EPI_ISL_651650\|202 | ···················· | ···················· | ···················· | ···················· | NNNNNNNNNN·········· | ···················· | 1 |
| S | 8529 | hCoV-19/England/CAMB-1B883B/2020\|EPI_ISL_651663\|20 | ···················· | NNNNNNNNNNNNNNNNNNNN | ···················· | ···················· | ···················· | ···················· | 1 |
| S | 8531 | hCoV-19/England/CAMB-1B8886/2020\|EPI_ISL_651665\|20 | ···················· | NNNNNNNNNNNNNNNNNNNN | ···················· | ···················· | ···················· | ···················· | 1 |
| S | 8548 | hCoV-19/England/CAMB-1B8C3F/2020\|EPI_ISL_651682\|20 | ···················· | NNNNNNNNNNNNNNNNNNNN | ···················· | ···················· | ···················· | ···················· | 1 |
| S | 8788 | hCoV-19/England/ALDP-B637A1/2020\|EPI_ISL_656470\|20 | ···················· | NNNNNNNNNNNNNNNNNNNN | ···················· | ···················· | ·T·················· | ···················· | 1 |
| T | 180 | hCoV-19/USA/CA-CZB-13134/2020\|EPI_ISL_672402\|2020- | ···················· | ·A·················· | ···················· | ···················· | NNNNNNNNNNNNNNNNNNNN | NNNNNNNNNNNNNNNNNNNN | 1 |
| T | 185 | hCoV-19/USA/CA-CZB-13129/2020\|EPI_ISL_672407\|2020- | ···················· | ·A·················· | ···················· | ···················· | NNNNNNNNNNNNNNNNNNNN | ···················· | 1 |
| T | 198 | hCoV-19/USA/CA-CZB-13113/2020\|EPI_ISL_672420\|2020- | ···················· | ·A·················· | ···················· | ···················· | ·NNNNNNNNNNNNNNNNNNN | NNNNNNNNNNNNNNNNNNNN | 1 |
| T | 201 | hCoV-19/USA/CA-CZB-12878/2020\|EPI_ISL_672423\|2020- | ···················· | ·A·················· | ···················· | ···················· | ···················· | ········NNNNNN······ | 1 |
| T | 202 | hCoV-19/USA/CA-CZB-12879/2020\|EPI_ISL_672424\|2020- | ···················· | ·A·················· | ···················· | ···················· | NNNNNNNNNNNNNNNNNNNN | NNNNNNNNNNNNNNNNNNNN | 1 |
| T | 205 | hCoV-19/USA/CA-CZB-13107/2020\|EPI_ISL_672427\|2020- | ···················· | ·A·················· | ···················· | ···················· | NNNNNNNNNNNNNNNNNNNN | ···················· | 1 |
| T | 207 | hCoV-19/USA/CA-CZB-13271/2020\|EPI_ISL_672429\|2020- | ···················· | ·A·················· | ···················· | ···················· | NNNNNNNNNNNNNNNNNNNN | NNNNNNNNNNNNNNNNNNNN | 1 |
| T | 208 | hCoV-19/USA/CA-CZB-13272/2020\|EPI_ISL_672430\|2020- | ···················· | ···················· | ···················· | ···················· | NNNNNNNNNNNN········ | NNNNNNNNNNNNNNNNNNNN | 1 |
| T | 210 | hCoV-19/USA/CA-CZB-13284/2020\|EPI_ISL_672432\|2020- | ···················· | ·A·················· | ···················· | ···················· | NNNNNNNNNNNNNNNNN··· | ···················· | 1 |
| T | 213 | hCoV-19/USA/CA-CZB-13304/2020\|EPI_ISL_672435\|2020- | ···················· | ·A·················· | ···················· | ···················· | NNNNNNNNNNNNNNNNNNNN | NNNNNNNNNNNNNNNNNNNN | 1 |
| T | 219 | hCoV-19/USA/CA-CZB-13343/2020\|EPI_ISL_672441\|2020- | ···················· | ···················· | ···················· | ···················· | ··NN···NNNNNNNNNNNN· | NNNNNNNNN········M·· | 1 |
| T | 220 | hCoV-19/USA/CA-CZB-13345/2020\|EPI_ISL_672442\|2020- | ···················· | ···················· | ···········T········ | ···················· | NNN················· | NNNNNNNNNNNNNNNNNNNN | 1 |
| T | 222 | hCoV-19/USA/CA-CZB-13349/2020\|EPI_ISL_672444\|2020- | ···················· | ···················· | ···················· | ···················· | NNNNNNNNNNNNNNNNNNNN | ····W··············· | 1 |
| T | 226 | hCoV-19/USA/CA-CZB-13360/2020\|EPI_ISL_672448\|2020- | ···················· | ···················· | ···················· | ···················· | NNNNNNNNNNNNNNNNNNNN | NNNNNNNNNNNNNNNNNNNN | 1 |
| T | 229 | hCoV-19/USA/CA-CZB-13374/2020\|EPI_ISL_672451\|2020- | ···················· | ···················· | ···················· | ···················· | NNNNNNNNNNNNNNNNNNNN | NNNNNNNNNNNNNNNNNNNN | 1 |
| T | 231 | hCoV-19/USA/CA-CZB-13377/2020\|EPI_ISL_672453\|2020- | ···················· | ···················· | ···················· | ···················· | NNNNNNN············· | ···················· | 1 |
| T | 242 | hCoV-19/USA/CA-CZB-13246/2020\|EPI_ISL_672485\|2020- | ···················· | ·A·················· | ···················· | ···················· | NNNN················ | ···················· | 1 |
| T | 250 | hCoV-19/USA/CA-CZB-13547/2020\|EPI_ISL_672493\|2020- | ···················· | ···················· | ···················· | ···················· | NNNNNNNNNNNNNNNNNNNN | ········NNNNNN······ | 1 |
| T | 284 | hCoV-19/England/LOND-12504E2/2020\|EPI_ISL_665214\|2 | ···················· | ···················· | ···················· | ···················· | NNNNNNNNNNNNNNNNNNNN | NNNNNNNNNNNNNNNNNNNN | 1 |
| T | 285 | hCoV-19/England/LOND-1250549/2020\|EPI_ISL_665215\|2 | ···················· | ···················· | ···················· | ···················· | ·NNNNNNNNNN········· | ···················· | 1 |
| T | 298 | hCoV-19/England/LOND-1250707/2020\|EPI_ISL_665228\|2 | ···················· | ···················· | ···················· | ···················· | NNNNNNNNNNNNNNNNNNNN | NNNNNNNNNNNNNNNNNNNN | 1 |
| T | 307 | hCoV-19/England/LOND-125089B/2020\|EPI_ISL_665237\|2 | ···················· | ···················· | ···················· | ···················· | NNNNNNNNNNNNNNNNNNNN | ···················· | 1 |
| T | 308 | hCoV-19/England/LOND-12508C8/2020\|EPI_ISL_665238\|2 | ···················· | ···················· | ···················· | ···················· | NNNNNNNNNNNNNNNNNNNN | ···················· | 1 |
| T | 314 | hCoV-19/England/LOND-1250AC2/2020\|EPI_ISL_665244\|2 | ···················· | ···················· | ···················· | ···················· | ·NNNNNNNNNNNNNNNNNNN | ···················· | 1 |
| T | 320 | hCoV-19/England/LOND-1250B65/2020\|EPI_ISL_665250\|2 | ···················· | ···················· | ···················· | ···················· | NNNNNNNNNNNNNNNNNNNN | ···················· | 1 |
| T | 325 | hCoV-19/Wales/PHWC-48CABF/2020\|EPI_ISL_666503\|2020 | ···················· | ···················· | ···················· | ···················· | NNNNNNN············· | ···················· | 1 |
| T | 335 | hCoV-19/Wales/PHWC-48CB70/2020\|EPI_ISL_666513\|2020 | ···················· | ···················· | ···················· | ···················· | NNNN················ | ········NN·········· | 1 |
| T | 351 | hCoV-19/Wales/PHWC-48CCF5/2020\|EPI_ISL_666529\|2020 | ···················· | ···················· | ···················· | ···················· | NNNNNNNNNNNNNNNNNNNN | NN······NNNNNN······ | 1 |
| T | 356 | hCoV-19/Wales/PHWC-48CD4D/2020\|EPI_ISL_666534\|2020 | ···················· | ···················· | ···················· | ···················· | NNNNNNNNNNNNNNNNNNNN | NN·····NNNNNNNNNNNNN | 1 |
| T | 494 | hCoV-19/USA/CA-CZB-13392/2020\|EPI_ISL_672456\|2020- | ···················· | ···················· | ···················· | ···················· | ·NNNNNNNNNNN········ | ···················· | 1 |
| T | 496 | hCoV-19/USA/CA-CZB-13397/2020\|EPI_ISL_672458\|2020- | ············T······· | ···················· | ···················· | ···················· | NNNNNNNNNNNNNNNNNNNN | NNNNNNNNNNNNNNNNNNNN | 1 |
| T | 498 | hCoV-19/USA/CA-CZB-13403/2020\|EPI_ISL_672460\|2020- | ···················· | ···················· | ···················· | ···················· | NNNNNNNNNNNNNNNNNNNN | NNNNNNNNNNNNNNNNNNNN | 1 |
| T | 499 | hCoV-19/USA/CA-CZB-13407/2020\|EPI_ISL_672461\|2020- | ···················· | ·A·················· | ···················· | ···················· | NNNN··········K····· | ···················· | 1 |
| T | 505 | hCoV-19/USA/CA-CZB-13421/2020\|EPI_ISL_672467\|2020- | ···················· | ·A·················· | ···················· | ···················· | NNNNNNNNNNNN········ | ···················· | 1 |
| T | 511 | hCoV-19/USA/CA-CZB-13454/2020\|EPI_ISL_672474\|2020- | ···················· | ···················· | ···················· | ···················· | NNNNNNNNNNNNNNNNNNNN | NNNNNNNNNNNNNNNNNNNN | 1 |
| T | 636 | hCoV-19/England/LOND-1250BCF/2020\|EPI_ISL_665253\|2 | ···················· | ···················· | ···················· | ···················· | NNNNNNNNNNNNNNNNNNNN | NNNNNNNNNNNNNNNNNNNN | 1 |
| T | 716 | hCoV-19/USA/CA-CZB-13018/2020\|EPI_ISL_672303\|2020- | ···················· | ·A·················· | ···················· | ···················· | N··················· | NNNNNNNNNNNNNNNNN··N | 1 |
| T | 718 | hCoV-19/USA/CA-CZB-13021/2020\|EPI_ISL_672305\|2020- | ···················· | ···················· | ···················· | ···················· | NNNNNNNNNNNNNNNNNNNN | NNNNNNNNNNNNNNNNNNNN | 1 |
| T | 719 | hCoV-19/USA/CA-CZB-13022/2020\|EPI_ISL_672306\|2020- | ···················· | ···················· | ···················· | ···················· | ···················· | ··NNN·········N····· | 1 |
| T | 725 | hCoV-19/USA/CA-CZB-13029/2020\|EPI_ISL_672312\|2020- | ···················· | ···················· | ···················· | ···················· | NNNNNNNNNNNN········ | NNNNNNNNNNNNNNNNNNNN | 1 |
| T | 727 | hCoV-19/USA/CA-CZB-13034/2020\|EPI_ISL_672314\|2020- | ···················· | ···················· | ···················· | ···················· | NNNNNNNNNNNNNNNNNNNN | NNNNNNNNNNNNNNNNNNNN | 1 |
| T | 730 | hCoV-19/USA/CA-CZB-13037/2020\|EPI_ISL_672317\|2020- | ···················· | ···················· | ···················· | ···················· | NNNNNNNNNNNNNNNNNNNN | NNNNNNNNNNNNNNNNNNNN | 1 |
| T | 741 | hCoV-19/USA/CA-CZB-13058/2020\|EPI_ISL_672328\|2020- | ···················· | ···················· | ···················· | ···················· | NNNNNNNNNNNNNNNNNNNN | NNNNNNNNNNNNNNNNNNNN | 1 |
| T | 744 | hCoV-19/USA/CA-CZB-13061/2020\|EPI_ISL_672331\|2020- | ···················· | ···················· | ···················· | ···················· | NNNNNNNNNNNNNNNNNNNN | NNNNNNNNNNNNNNNNNNNN | 1 |
| T | 745 | hCoV-19/USA/CA-CZB-13062/2020\|EPI_ISL_672332\|2020- | ···················· | ···················· | ···················· | ···················· | NNNN················ | ·················M·· | 1 |
| T | 758 | hCoV-19/USA/CA-CZB-5707/2020\|EPI_ISL_672389\|2020-0 | ···················· | ·A·················· | ···················· | ···················· | NNNNNNNNNNNNNNNNNNNN | ···················· | 1 |
| T | 759 | hCoV-19/USA/CA-CZB-5706/2020\|EPI_ISL_672390\|2020-0 | ···················· | ···················· | ···················· | ···················· | NNNNNNNNNNNNNNNNNNNN | NNNNNNNNNNNNNNNNNNNN | 1 |
| T | 761 | hCoV-19/USA/CA-CZB-5684/2020\|EPI_ISL_672392\|2020-0 | ···················· | ···················· | ···················· | ···················· | NNNNNNNNNNNNNNNNNNNN | ···················· | 1 |
| T | 841 | hCoV-19/Wales/PHWC-48C457/2020\|EPI_ISL_666430\|2020 | ···················· | ···················· | ···················· | ···················· | NNNNNNNNNNNNNNNNNNNN | ···················· | 1 |
| T | 842 | hCoV-19/Wales/PHWC-48C475/2020\|EPI_ISL_666431\|2020 | ···················· | ···················· | ···················· | ···················· | NNNN················ | ···················· | 1 |
| T | 852 | hCoV-19/Wales/PHWC-48C563/2020\|EPI_ISL_666441\|2020 | ···················· | ···················· | ···················· | ···················· | NNNNNNNNNNNNNN······ | ···················· | 1 |
| T | 863 | hCoV-19/Wales/PHWC-48C660/2020\|EPI_ISL_666452\|2020 | ···················· | ···················· | ···················· | ···················· | NNNNNNNNNNNNNNNNNNNN | ···················· | 1 |
| T | 990 | hCoV-19/USA/CA-CZB-13080/2020\|EPI_ISL_672342\|2020- | ···················· | ···················· | ···················· | ···················· | NNNNNNNNNNNNNNNNNNNN | NNNNNNNNNNNNNNNNNNNN | 1 |
| T | 991 | hCoV-19/USA/CA-CZB-13083/2020\|EPI_ISL_672343\|2020- | ···················· | ···················· | ···················· | ···················· | NNNNNNNNNNNNNNNNNNNN | NNNNNNNNNNNNNNNNNNNN | 1 |
| T | 992 | hCoV-19/USA/CA-CZB-13086/2020\|EPI_ISL_672344\|2020- | ···················· | ···················· | ···················· | ···················· | NNNNNNNNNNNNNNNNNNNN | NNNNNNNNNNNNNNNNNNNN | 1 |
| T | 994 | hCoV-19/USA/CA-CZB-13089/2020\|EPI_ISL_672346\|2020- | ···················· | ···················· | ···················· | ···················· | NNNNNNNNNNNNNNNNNNNN | NNNNNNNNNNNNNNNNNNNN | 1 |
| T | 996 | hCoV-19/USA/CA-CZB-13094/2020\|EPI_ISL_672348\|2020- | ···················· | ···················· | ···················· | ···················· | NNNNNNNNNNNNNNNNNNNN | NNNNNNNNNNNNNNNNNNNN | 1 |
| T | 998 | hCoV-19/USA/CA-CZB-13096/2020\|EPI_ISL_672350\|2020- | ···················· | ···················· | ···················· | ···················· | NNNNNNNNNNNNNNNNNNNN | ······NNNNNNNNNNNNNN | 1 |
| T | 1003 | hCoV-19/USA/CA-CZB-5773/2020\|EPI_ISL_672355\|2020-0 | ···················· | ···················· | ···················· | ···················· | NNNN················ | ···················· | 1 |
| T | 1117 | hCoV-19/England/QEUH-AF4D3B/2020\|EPI_ISL_662894\|20 | ···················· | NNNNNNNNNNNNNNNNNNNN | ···················· | ···················· | ···················· | ···················· | 1 |
| T | 1153 | hCoV-19/England/LIVE-DBD57F/2020\|EPI_ISL_665134\|20 | ···················· | ···················· | ···················· | ···················· | ·······NNNNNNNNNNNNN | ···················· | 1 |
| T | 1155 | hCoV-19/England/LIVE-DBE8DF/2020\|EPI_ISL_665136\|20 | ··················N· | ···················· | ···················· | ···················· | NNNNNNNNNNNNNNNNNNNN | NNNNNNNNNNNNNNNNNNNN | 1 |
| T | 1156 | hCoV-19/England/LIVE-DBE8FD/2020\|EPI_ISL_665137\|20 | ···················· | ···················· | ···················· | ···················· | NNNNNNNNNNNNNNNNNNNN | NNNNNNNNNNNNNNNNNNNN | 1 |
| T | 1157 | hCoV-19/England/LIVE-DBE972/2020\|EPI_ISL_665138\|20 | ···················· | ···················· | ···················· | ···················· | NNNNNNNNNNNNNNNNNNNN | NNNNNNNNNNNNNNNNNNNN | 1 |
| T | 1158 | hCoV-19/England/LIVE-DBEABB/2020\|EPI_ISL_665139\|20 | ···················· | ···················· | ···················· | ···················· | NNNNNNNNNNNNNNNNNNNN | NNNNNNNNNNNNNNNNNNNN | 1 |

| | | | | | | | | |
|---|---|---|---|---|---|---|---|---|
| T | 1160 hCoV-19/England/LIVE-DC1262/2020\|EPI_ISL_665141\|20 | ···················· | ·········T·········· | ···················· | ···················· | NNNNNNNNNNNNNNNNNNNN | NNNNNNNNNNNNNNNNNNNN | 1 |
| T | 1161 hCoV-19/England/LIVE-DC12EA/2020\|EPI_ISL_665142\|20 | ···················· | ···················· | ···················· | ···················· | NNNNNNNNNNNNNNNNNNNN | NNNNNNNNNNNNNNNNNNNN | 1 |
| T | 1165 hCoV-19/England/LOND-124F1FE/2020\|EPI_ISL_665146\|2 | ···················· | ···················· | ···················· | ···················· | NNNNNNNNNNNN········ | ···················· | 1 |
| T | 1166 hCoV-19/England/LOND-124F246/2020\|EPI_ISL_665147\|2 | ···················· | ···················· | ···················· | ···················· | NNNNNNNNNNNNNNNNNNNN | NNNNNNNNNNNNNNN··NNN | 1 |
| T | 1169 hCoV-19/England/LOND-124F2FB/2020\|EPI_ISL_665150\|2 | ···················· | ···················· | ···················· | ···················· | NNNNNNNNNNNNNNNNNNNN | NNNNNNNNNNNNNNNNNNNN | 1 |
| T | 1180 hCoV-19/England/LOND-124F5D4/2020\|EPI_ISL_665161\|2 | ···················· | ···················· | ···················· | ···················· | NNNNNNNNNNNNN······· | ···················· | 1 |
| T | 1191 hCoV-19/England/LOND-124F7A1/2020\|EPI_ISL_665172\|2 | ···················· | ···················· | ···················· | ···················· | NNN··········NNNNNNN | ···················· | 1 |
| T | 1193 hCoV-19/England/LOND-124F835/2020\|EPI_ISL_665174\|2 | ···················· | ···················· | ···················· | ···················· | ·NN·NNNNNNNNN······· | ···················· | 1 |
| T | 1194 hCoV-19/England/LOND-124F862/2020\|EPI_ISL_665175\|2 | ···················· | ···················· | ···················· | ···················· | NNNN···NNNNNNNNNNNNN | ···················· | 1 |
| T | 1196 hCoV-19/England/LOND-124F8CC/2020\|EPI_ISL_665177\|2 | ···················· | ···················· | ···················· | ···················· | NNNNNNNNNNNNNNNNNNNN | ···················· | 1 |
| T | 1198 hCoV-19/England/LOND-124F941/2020\|EPI_ISL_665179\|2 | ···················· | ···················· | ···················· | ···················· | NNNNNNNNNNNN········ | NNNNNNNN············ | 1 |
| T | 1219 hCoV-19/Wales/PHWC-48C6F7/2020\|EPI_ISL_666459\|2020 | ···················· | ···················· | ···················· | ···················· | NNNNNNNNNNNNNNNNNNNN | NNNNNNNNNNNNNNNNNNNN | 1 |
| T | 1239 hCoV-19/Wales/PHWC-48C888/2020\|EPI_ISL_666479\|2020 | ···················· | ···················· | ···················· | ···················· | ···················· | NNNN················ | 1 |
| T | 1255 hCoV-19/Wales/PHWC-48CA19/2020\|EPI_ISL_666495\|2020 | ···················· | ···················· | ···················· | ···················· | NNNNNNNNNNNN········ | ···················· | 1 |
| T | 1449 hCoV-19/USA/CA-CZB-10097/2020\|EPI_ISL_672212\|2020- | ···················· | ···················· | ···················· | ···················· | ···················· | NNNNNNNNNNNNNNN····· | 1 |
| T | 1453 hCoV-19/USA/CA-CZB-10108/2020\|EPI_ISL_672216\|2020- | ···················· | ···················· | ···················· | ···················· | NNNNNNNNNNNNNN······ | ···················· | 1 |
| T | 1459 hCoV-19/USA/CA-CZB-10123/2020\|EPI_ISL_672222\|2020- | ···················· | ···················· | ···················· | ···················· | ·NNNNNNNNNN········· | NNNNNNNNNNNNNNN····· | 1 |
| T | 1472 hCoV-19/USA/CA-CZB-10156/2020\|EPI_ISL_672235\|2020- | ···················· | ·A·················· | ···················· | ···················· | NNNNNNNNNNNNNNNNNNNN | NNNNNNNNNNNNNNNNNNNN | 1 |
| T | 1483 hCoV-19/USA/CA-CZB-10178/2020\|EPI_ISL_672246\|2020- | ···················· | ···················· | ···················· | ···················· | NNNNNNNNNNNNNNNNNNNN | NNNNNNNNNNNNNNNNNNNN | 1 |
| T | 1493 hCoV-19/USA/CA-CZB-10198/2020\|EPI_ISL_672256\|2020- | ···················· | ···················· | ···················· | ···················· | NNNNNNNNNNNNNNNNNNNN | NNNNNNNNNNNNNNNNNNNN | 1 |
| T | 1504 hCoV-19/USA/CA-CZB-10232/2020\|EPI_ISL_672267\|2020- | ···················· | ···················· | ···················· | ···················· | NNNNNNNNNNNNNNNNNNNN | NNNNNNNNNNNNNNNNNNNN | 1 |
| T | 1515 hCoV-19/USA/CA-CZB-11881/2020\|EPI_ISL_672278\|2020- | ···················· | ·A·················· | ···················· | ···················· | NNNNNNNNNNNNNNNNNNNN | NNNNNNNNNNNNNNN····· | 1 |
| T | 1525 hCoV-19/USA/CA-CZB-11912/2020\|EPI_ISL_672288\|2020- | ···················· | ·A·················· | ···················· | ···················· | NNNNNNNNNNNNNNNNNNNN | NNNNNNNNNNNNNNNNNNNN | 1 |
| T | 1527 hCoV-19/USA/CA-CZB-11915/2020\|EPI_ISL_672290\|2020- | ···················· | ·A·················· | ···················· | ···················· | NNNNNNNNNNNNNNNNNNNN | ···················· | 1 |
| T | 1569 hCoV-19/England/EXET-138EF5/2020\|EPI_ISL_665047\|20 | ···················· | ···················· | ···················· | ···················· | NNNNNNNNNNNNNNNNNNNN | NNNNNNNNNNNNNNNNNNNN | 1 |
| T | 1570 hCoV-19/England/EXET-138F01/2020\|EPI_ISL_665048\|20 | ···················· | ···················· | ···················· | ···················· | NNNNNNNNNNNNNNNNNNNN | NNNNNNNNNNNNNNNNNNNN | 1 |
| T | 1571 hCoV-19/England/EXET-138F2F/2020\|EPI_ISL_665049\|20 | ···················· | ···················· | ···················· | ···················· | NNNNNNNNNNNNNNNNNNNN | NNNNNNNNNNNNNNNNNNNN | 1 |
| T | 1572 hCoV-19/England/EXET-138F3E/2020\|EPI_ISL_665050\|20 | ···················· | ···················· | ···················· | ···················· | NNNNNNNNNNNNNNNNNNNN | NNNNNNNNNNNNNNNNNNNN | 1 |
| T | 1573 hCoV-19/England/EXET-138FB6/2020\|EPI_ISL_665051\|20 | ···················· | ···················· | ···················· | ···················· | NNNNNNNNNNNNNNNNNNNN | NNNNNNNNNNNNNNNNNNNN | 1 |
| T | 1574 hCoV-19/England/EXET-1390A3/2020\|EPI_ISL_665052\|20 | ···················· | ···················· | ···················· | ···················· | NNNNNNNNNNNNNNNNNNNN | NNNNNNNNNNNNNNNNNNNN | 1 |
| T | 1576 hCoV-19/England/EXET-139119/2020\|EPI_ISL_665054\|20 | ···················· | ···················· | ···················· | ···················· | NNNNNNNNNNNNNNNNNNNN | NNNNNNNNNNNNNNNNNNNN | 1 |
| T | 1577 hCoV-19/England/EXET-139164/2020\|EPI_ISL_665055\|20 | ···················· | ···················· | ···················· | ···················· | NNNNNNNNNNNNNNNNNNNN | NNNNNNNNNNNNNNNNNNNN | 1 |
| T | 1579 hCoV-19/England/EXET-139191/2020\|EPI_ISL_665057\|20 | ···················· | ···················· | ···················· | ···················· | NNNNNNNNNNNNNNNNNNNN | NNNNNNNNNNNNNNNNNNNN | 1 |
| T | 1580 hCoV-19/England/EXET-1391BF/2020\|EPI_ISL_665058\|20 | ···················· | ···················· | ···················· | ···················· | NNNNNNNNNNNNNNNNNNNN | NNNNNNNNNNNNNNNNNNNN | 1 |
| T | 1581 hCoV-19/England/EXET-13929E/2020\|EPI_ISL_665059\|20 | ···················· | ···················· | ···················· | ···················· | NNNNNNNNNNNNNNNNNNNN | NNNNNNNNNNNNNNNNNNNN | 1 |
| T | 1582 hCoV-19/England/EXET-1392DA/2020\|EPI_ISL_665060\|20 | ···················· | ···················· | ···················· | ···················· | NNNNNNNNNNNNNNNNNNNN | NNNNNNNNNNNNNNNNNNNN | 1 |
| T | 1583 hCoV-19/England/EXET-1392E9/2020\|EPI_ISL_665061\|20 | ···················· | ···················· | ···················· | ···················· | NNNNNNNNNNNNNNNNNNNN | NNNNNNNNNNNNNNNNNNNN | 1 |
| T | 1585 hCoV-19/England/EXET-13935F/2020\|EPI_ISL_665063\|20 | ···················· | ···················· | ···················· | ···················· | NNNNNNNNNNNNNNNNNNNN | NNNNNNNNNNNNNNNNNNNN | 1 |
| T | 1586 hCoV-19/England/EXET-13936E/2020\|EPI_ISL_665064\|20 | ···················· | ···················· | ···················· | ···················· | NNNNNNNNNNNNNNNNNNNN | NNNNNNNNNNNNNNNNNNNN | 1 |
| T | 1587 hCoV-19/England/EXET-13937D/2020\|EPI_ISL_665065\|20 | ···················· | ···················· | ···················· | ···················· | NNNNNNNNNNNNNNNNNNNN | NNNNNNNNNNNNNNNNNNNN | 1 |
| T | 1664 hCoV-19/Wales/PHWC-48C141/2020\|EPI_ISL_666398\|2020 | ···················· | ···················· | ···················· | ···················· | NNNNNNNNNNNNNNNNNNNN | NNNNNNNNNNNNNNNNNNNN | 1 |
| T | 1772 hCoV-19/USA/CA-CZB-9792/2020\|EPI_ISL_672179\|2020-0 | ···················· | ···················· | ···················· | ···················· | NNNNNNNNNNNNNNNNNNNN | ···················· | 1 |
| T | 1933 hCoV-19/England/CAMB-1BB5B9/2020\|EPI_ISL_665012\|20 | ···················· | ···················· | ···················· | ···················· | NNNNNNNNNNNNNNNNNNNN | NNNNNNNNNNNNNNNNNNNN | 1 |
| T | 1937 hCoV-19/England/CAMB-1BB65C/2020\|EPI_ISL_665016\|20 | ···················· | ···················· | ··············G····· | ···················· | NNNNNNNNNNNNNNNNNNNN | NNNNNNNNNNNNNNNNNNNN | 1 |
| T | 1981 hCoV-19/USA/CA-CZB-13470/2020\|EPI_ISL_672102\|2020- | ···················· | ·A·················· | ···················· | ···················· | NNNN················ | ··········R········· | 1 |
| T | 1989 hCoV-19/USA/CA-CZB-9526/2020\|EPI_ISL_672119\|2020-0 | ···················· | ···················· | ···················· | ···················· | NNN··NNNNNNNNNNNNNNN | ···················· | 1 |
| T | 1995 hCoV-19/USA/CA-CZB-9541/2020\|EPI_ISL_672125\|2020-0 | ···················· | ···················· | ···················· | ···················· | NNNNNNNNNNNNNNNNNNNN | NNNNNNNNNNNNNNNNNNNN | 1 |
| T | 1996 hCoV-19/USA/CA-CZB-9545/2020\|EPI_ISL_672126\|2020-0 | ···················· | ···················· | ···················· | ···················· | NNNNNNNNNNNNNNNNNNNN | NNNNNNNNNNNNNNNNNNNN | 1 |
| T | 2026 hCoV-19/USA/CA-CZB-9707/2020\|EPI_ISL_672156\|2020-0 | ···················· | ···················· | ···················· | ···················· | NNNNNNNNNNNNNNNNNNNN | NNNNNNNNNNNNNNNNNNNN | 1 |
| T | 2027 hCoV-19/USA/CA-CZB-9714/2020\|EPI_ISL_672157\|2020-0 | ···················· | ···················· | ···················· | ···················· | ···················· | NNNNN·······NNNNNNNN | 1 |
| T | 2045 hCoV-19/USA/CA-CZB-9776/2020\|EPI_ISL_672175\|2020-0 | ···················· | ·A·················· | ···················· | ···················· | NNNNNNNNNNNNNNNNNNNN | NNNNNNNNNNNNNNNNNNNN | 1 |
| T | 2057 hCoV-19/Wales/PHWC-48B19D/2020\|EPI_ISL_666201\|2020 | ···················· | ···················· | ···················· | ···················· | NNNNNNNNNNNNNNNNNNNN | NNNNNNNNNNNNNNNNNNNN | 1 |
| T | 2063 hCoV-19/Wales/PHWC-48B1F7/2020\|EPI_ISL_666207\|2020 | ···················· | ···················· | ···················· | ···················· | NNNNNNNNNNNN········ | ···················· | 1 |
| T | 2087 hCoV-19/Wales/PHWC-48B3C4/2020\|EPI_ISL_666231\|2020 | ···················· | ···················· | ···················· | ···················· | NNNNNNNNNNNNNNNNNNNN | ·················NNN | 1 |
| T | 2113 hCoV-19/Wales/PHWC-48B5CE/2020\|EPI_ISL_666257\|2020 | ···················· | ···················· | ···················· | ···················· | NNNNNNNNNNNNNNNNNNNN | ···················· | 1 |
| T | 2302 hCoV-19/USA/CA-CZB-13497/2020\|EPI_ISL_672099\|2020- | ···················· | ·A·················· | ···················· | ···················· | N··················· | NNNNN··············· | 1 |
| T | 2318 hCoV-19/England/MILK-A698F4/2020\|EPI_ISL_665760\|20 | ···················· | ···················· | ···················· | ···················· | NNNNNNNNNNNNNNNNNNNN | NNNNNNNNNNNNNNNNNNNN | 1 |
| T | 2494 hCoV-19/Canada/ON-PHL-20-02419/2020\|EPI_ISL_671531 | ···················· | ···················· | ···················· | ···················· | NNNNNNNNNNNNNNNNNNNN | NNNNNNNNNNNNNNNNNNNN | 1 |
| T | 2495 hCoV-19/Canada/ON-PHL-20-02433/2020\|EPI_ISL_671532 | ···················· | ···················· | ···················· | ···················· | NNNN················ | ···················· | 1 |
| T | 2499 hCoV-19/Canada/ON-PHL-20-02426/2020\|EPI_ISL_671536 | ···················· | ···················· | ···················· | ···················· | NNNNNNNNNNNNNNNNNNNN | NNNNNNNNNNNNNNNN···· | 1 |
| T | 2500 hCoV-19/Canada/ON-PHL-20-02394/2020\|EPI_ISL_671537 | ···················· | ···················· | ···················· | ···················· | NNNN················ | ···················· | 1 |
| T | 2501 hCoV-19/Canada/ON-PHL-20-02410/2020\|EPI_ISL_671538 | ···················· | ···················· | ···················· | ···················· | NNNNNNNNNNNNNNNNNNNN | NNNNNNNNNNNNNNNNNNNN | 1 |
| T | 2502 hCoV-19/Canada/ON-PHL-20-02409/2020\|EPI_ISL_671539 | ···················· | ···················· | ···················· | ···················· | NNNNNNNNNNNNNNNNNNNN | NNNNNNNNN··········· | 1 |
| T | 2503 hCoV-19/Canada/ON-PHL-20-02412/2020\|EPI_ISL_671540 | ···················· | ···················· | ···················· | ···················· | NNNNNNNNNNNNNNNNNNNN | ···················· | 1 |
| T | 2504 hCoV-19/Canada/ON-PHL-20-02411/2020\|EPI_ISL_671541 | ···················· | ···················· | ···················· | ···················· | NNNNNNNNNNNNNNNNNNNN | NNNNNNNNNNNNNNNNNNNN | 1 |
| T | 2505 hCoV-19/Canada/ON-PHL-20-02449/2020\|EPI_ISL_671542 | ···················· | ···················· | ···················· | ···················· | NNNNNNNNNNNNNNNNNNNN | NNN················· | 1 |
| T | 2513 hCoV-19/Canada/ON-PHL-20-02306/2020\|EPI_ISL_671550 | ···················· | ···A················ | ···················· | ···················· | NNNNNNNNNNNNNNNNNNNN | NNNNNNNNNNNNNNNNNNNN | 1 |
| T | 2522 hCoV-19/Canada/ON-PHL-20-02271/2020\|EPI_ISL_671559 | ···················· | ···················· | ···················· | ···················· | NNNNNNNNNNNNNNNNNNNN | NNNNNNNNNNNNNNNNNNNN | 1 |
| T | 2523 hCoV-19/Canada/ON-PHL-20-02272/2020\|EPI_ISL_671560 | ···················· | ···················· | ···················· | ···················· | NNNNNNNNNNNNNNNNNNNN | NNNNNNNNNNNNNNNNNNNN | 1 |
| T | 2530 hCoV-19/Canada/ON-PHL-20-02335/2020\|EPI_ISL_671567 | ···················· | ···················· | ···················· | ···················· | NNNNNNNNNNNNNNNNNNNN | ···················· | 1 |
| T | 2540 hCoV-19/Canada/ON-PHL-20-02440/2020\|EPI_ISL_671577 | ···················· | ···················· | ···················· | ···················· | NNNN················ | ···················· | 1 |
| T | 2541 hCoV-19/Canada/ON-PHL-20-02441/2020\|EPI_ISL_671578 | ···················· | ···················· | ··········T········· | ···················· | NNNNNNNNNNNNNNNNNNNN | ···················· | 1 |
| T | 2545 hCoV-19/Canada/ON-PHL-20-02438/2020\|EPI_ISL_671582 | ···················· | ···················· | ···················· | ···················· | ···················· | NNNNNNNNNNNNNNN····· | 1 |
| T | 2549 hCoV-19/Canada/ON-PHL-20-02478/2020\|EPI_ISL_671586 | ···················· | ···················· | ···················· | ···················· | NNNN················ | ···················· | 1 |
| T | 2550 hCoV-19/Canada/ON-PHL-20-02475/2020\|EPI_ISL_671587 | ···················· | ···················· | ···················· | ···················· | NNNN················ | ···················· | 1 |
| T | 2553 hCoV-19/Canada/ON-PHL-20-02408/2020\|EPI_ISL_671590 | ···················· | ···················· | ···················· | ···················· | NNNNNNNNNNNNNNNNNNNN | NNNNNNNNNNNNNNNNNNNN | 1 |
| T | 2554 hCoV-19/Canada/ON-PHL-20-02413/2020\|EPI_ISL_671591 | ···················· | ···················· | ···················· | ···················· | NNNNNNNNNNNNNNNNNNNN | NNNNNNNNNNNNNNNNNNNN | 1 |
| T | 2555 hCoV-19/Canada/ON-PHL-20-02422/2020\|EPI_ISL_671592 | ···················· | ···················· | ···················· | ···················· | NNNNNNNNNNNNNNNNNNNN | ···················· | 1 |
| T | 2557 hCoV-19/Canada/ON-PHL-20-02424/2020\|EPI_ISL_671594 | ···················· | ···················· | ···················· | ···················· | NNNNNNNNNNNNNNNNNNNN | NNNNNNNNNNNNNNNNNNNN | 1 |
| T | 2558 hCoV-19/Canada/ON-PHL-20-02226/2020\|EPI_ISL_671595 | ···················· | ···················· | ···················· | ···················· | NNNNNNNNNNNNNNNNNNNN | ···················· | 1 |

| | | | | | | | | |
|---|---|---|---|---|---|---|---|---|
| T | 2559 hCoV-19/Canada/ON-PHL-20-02406/2020\|EPI_ISL_671596 | ···················· | ···················· | ···················· | ···················· | NNNN················ | ···················· | 1 |
| T | 2638 hCoV-19/England/QEUH-AF3E66/2020\|EPI_ISL_663093\|20 | ···················· | ···················· | ···················· | NNNNNNNNNNNNNNNNNNNN | ···················· | ···················· | 1 |
| T | 2801 hCoV-19/Canada/ON-PHL-20-02233/2020\|EPI_ISL_671504 | ···················· | ···················· | ···················· | ···················· | NNNNNNNNNNNNNNNNNNNN | NNNNNNNNNNNNNNNNNNNN | 1 |
| T | 2804 hCoV-19/Canada/ON-PHL-20-02211/2020\|EPI_ISL_671507 | ···················· | ···A················ | ···················· | ···················· | ···················· | NNNNNNNNNNNNNNNN·NNN | 1 |
| T | 2808 hCoV-19/Canada/ON-PHL-20-02240/2020\|EPI_ISL_671511 | ···················· | ···A················ | ···················· | ···················· | NNNNNNNNNNNNNNNNNNNN | ···················· | 1 |
| T | 2816 hCoV-19/Canada/ON-PHL-20-02225/2020\|EPI_ISL_671519 | ···················· | ···A················ | ···················· | ···················· | NNNNNNNNNNNN········ | ···················· | 1 |
| T | 3027 hCoV-19/Hungary/SRC-2140/2020\|EPI_ISL_671417\|2020- | ···················· | ···················· | ···················· | ···················· | NNNNNNNNNNNNNNNNNNNN | NNNNNNNNNNNNNNNNNNNN | 1 |
| T | 3059 hCoV-19/Hungary/UD-93823/2020\|EPI_ISL_671486\|2020- | ···················· | ···················· | ···················· | ···················· | NNNNNNNNNNNNNNNNNNNN | NNNNNNNNNNNNNNNNNNNN | 1 |
| T | 3070 hCoV-19/Canada/ON-PHL-20-02197/2020\|EPI_ISL_671497 | ···················· | ···················· | ···················· | ···················· | NNNNNNNNNNNNNNNNNNNN | ···················· | 1 |
| T | 3125 hCoV-19/USA/FL-BPHL-2215/2020\|EPI_ISL_666837\|2020- | ···················· | ···················· | ···················· | ···················· | NNNNNNNNNNNNNNNNNNNN | ·······NNNNNNNNNNNNN | 1 |
| T | 3126 hCoV-19/USA/FL-BPHL-2218/2020\|EPI_ISL_666840\|2020- | ···················· | ···················· | ···················· | ···················· | NNNNNNNNNNNNNNNNNNNN | ···················· | 1 |
| T | 3127 hCoV-19/USA/FL-BPHL-2220/2020\|EPI_ISL_666842\|2020- | ···················· | ···················· | ···················· | ···················· | NNNNNNNNNNNNNNNNNNNN | NNNNN·······NNNNN·NN | 1 |
| T | 3634 hCoV-19/Australia/VIC17718/2020\|EPI_ISL_664008\|202 | ···················· | ···················· | ···················· | ···················· | NNNNNNNNNNNNNNNNNNNN | ···················· | 1 |
| T | 3635 hCoV-19/Australia/VIC17720/2020\|EPI_ISL_664009\|202 | ···················· | ···················· | ···················· | ···················· | NNNNNNNNNNNNNNNNNNNN | NNNNNNNNNNNNNNN··NNN | 1 |
| T | 3636 hCoV-19/Australia/VIC17721/2020\|EPI_ISL_664010\|202 | ···················· | ···················· | ···················· | ···················· | NNNNNNNNNNNNNNNNNNNN | ···················· | 1 |
| T | 3637 hCoV-19/Australia/VIC17722/2020\|EPI_ISL_664011\|202 | ···················· | ···················· | ···················· | ···················· | NNNNNNNNNNNNNNNNNNNN | ···················· | 1 |
| T | 3640 hCoV-19/Australia/VIC17725/2020\|EPI_ISL_664014\|202 | ···················· | ···················· | ···················· | ···················· | NNNNNNNNNNNNNNNNNNNN | ·············S······ | 1 |
| T | 3641 hCoV-19/Australia/VIC17726/2020\|EPI_ISL_664015\|202 | ···················· | ···················· | ···················· | ···················· | NNNNNNNNNNNNNNNNNNNN | NNNNNNNNNNNNNNNNNNNN | 1 |
| T | 3642 hCoV-19/Australia/VIC17727/2020\|EPI_ISL_664016\|202 | ···················· | ···················· | ···················· | ···················· | NNNNNNNNNNNNNNNNNNNN | ···················· | 1 |
| T | 3664 hCoV-19/France/ARA-SC056/2020\|EPI_ISL_666670\|2020- | ···················· | ···················· | ···················· | ···················· | NNNNNNNNNNNN········ | NN·················· | 1 |
| T | 3668 hCoV-19/France/ARA-SC062/2020\|EPI_ISL_666674\|2020- | ···················· | ···················· | ···················· | ···················· | NNNNNNNNNNNNNNNNNNNN | ···················· | 1 |
| T | 3699 hCoV-19/France/NAQ-SC295/2020\|EPI_ISL_666716\|2020- | ···················· | ···················· | ···················· | ···················· | NNNNNNNNNNNNNNNNNNNN | ···················· | 1 |
| T | 3806 hCoV-19/USA/CA-CZB-13537/2020\|EPI_ISL_672500\|2020- | ···················· | ···················· | ···················· | ···················· | NNNNNNNNNNNNNNNNNNNN | NNNNNNNNNNNNNNNNNNNN | 1 |
| T | 3809 hCoV-19/USA/CA-CZB-13557/2020\|EPI_ISL_672503\|2020- | ···················· | ·A·················· | ···················· | ···················· | NNNNNNNNNNNNNNNNNNNN | NNNNNNNNNNNNNNNNNNNN | 1 |
| T | 3813 hCoV-19/USA/CA-CZB-13524/2020\|EPI_ISL_672507\|2020- | ···················· | ·A·················· | ···················· | ···················· | NNNNNNNNNNNNNNNNNNNN | NNNNNNNNNNNNNNNNNNNN | 1 |
| T | 3820 hCoV-19/USA/CA-CZB-13510/2020\|EPI_ISL_672514\|2020- | ···················· | ·A·················· | ···················· | ···················· | NNNNNNNNNNNNNNNNNNNN | NNNNNNNNNNNNNNN····· | 1 |
| T | 3834 hCoV-19/USA/CA-CZB-13475/2020\|EPI_ISL_672528\|2020- | ···················· | ···················· | ···················· | ···················· | NNNNNNNNNNNNNNNNNNNN | NNNNNNNNNNNNNNNNNNNN | 1 |
| T | 3836 hCoV-19/USA/CA-CZB-13465/2020\|EPI_ISL_672530\|2020- | ···················· | ···················· | ···················· | ···················· | NNNNNNNNNNNNNNNNNNNN | ·············NN····· | 1 |
| T | 3909 hCoV-19/Australia/VIC16633/2020\|EPI_ISL_663546\|202 | ···················· | ···················· | ···················· | ···················· | NNNNNNNNNNNNNNNNNNNN | ···················· | 1 |
| T | 3912 hCoV-19/Australia/VIC16620/2020\|EPI_ISL_663549\|202 | ···················· | ···················· | ···················· | ···················· | NNNNNNNNNNNNNNNNNNNN | NNNNNNNNNNNNNNNNNNNN | 1 |
| T | 3913 hCoV-19/Australia/VIC16660/2020\|EPI_ISL_663550\|202 | ···················· | ···················· | ···················· | ···················· | NNNNNNNNNNNNNNNNN··· | NNNNNNNNNNNNNNNNNNNN | 1 |
| T | 3914 hCoV-19/Australia/VIC16722/2020\|EPI_ISL_663551\|202 | ···················· | ···················· | ···················· | ···················· | NNNNNNNNNNNNNNNNNNNN | ···················· | 1 |
| T | 3915 hCoV-19/Australia/VIC16725/2020\|EPI_ISL_663552\|202 | ···················· | ···················· | ···················· | ···················· | NNNNNNNNNNNNNNNNNNNN | NNNNNNNNNNNNNNNNNNNN | 1 |
| T | 3916 hCoV-19/Australia/VIC16729/2020\|EPI_ISL_663555\|202 | ···················· | ···················· | ···················· | ···················· | NNNN················ | ···················· | 1 |
| T | 3921 hCoV-19/Australia/VIC16739/2020\|EPI_ISL_663560\|202 | ···················· | ···················· | ···················· | ···················· | NNNNNNNNNNNNNNNN···· | ···················· | 1 |
| T | 3922 hCoV-19/Australia/VIC16741/2020\|EPI_ISL_663561\|202 | ···················· | ·····R·············· | ···················· | ···················· | ··N················· | NNNNN··············· | 1 |
| T | 3924 hCoV-19/Australia/VIC16743/2020\|EPI_ISL_663563\|202 | ···················· | ···················· | ···················· | ···················· | NNNNNNNNNNNN········ | ···················· | 1 |
| T | 3925 hCoV-19/Australia/VIC16745/2020\|EPI_ISL_663564\|202 | ···················· | ···················· | ···················· | ···················· | N··········S········ | NNNN················ | 1 |
| T | 3926 hCoV-19/Australia/VIC16746/2020\|EPI_ISL_663565\|202 | ···················· | ···················· | ···················· | ···················· | NNNNNNNNNNNNNNNNNNNN | NN·················· | 1 |
| T | 3927 hCoV-19/Australia/VIC16747/2020\|EPI_ISL_663566\|202 | ···················· | ···················· | ···················· | ···················· | NNNNNNNNNNNNNNNNNNNN | ···················· | 1 |
| T | 3928 hCoV-19/Australia/VIC16750/2020\|EPI_ISL_663567\|202 | ···················· | ···················· | ···················· | ···················· | NNNNNNNNNNNNNNNNNNNN | NNNNNNNNNNNNNNNNNNNN | 1 |
| T | 3930 hCoV-19/Australia/VIC16754/2020\|EPI_ISL_663569\|202 | ···················· | ···················· | ···················· | ···················· | NNNNNNNNNNNNNNNNNNNN | ···················· | 1 |
| T | 3931 hCoV-19/Australia/VIC16755/2020\|EPI_ISL_663570\|202 | ···················· | ···················· | ···················· | ···················· | NNNNNNNNNNNNNNNNNNNN | NNNNNNNNN··········· | 1 |
| T | 3932 hCoV-19/Australia/VIC16759/2020\|EPI_ISL_663571\|202 | ···················· | ···················· | ···················· | ···················· | NNNNNNNNNNNNNNNNNNNN | ···················· | 1 |
| T | 3944 hCoV-19/Australia/VIC16798/2020\|EPI_ISL_663583\|202 | ···················· | ···················· | ···················· | ···················· | NNNNNNNNNNNNNNNNNNNN | ···················· | 1 |
| T | 3961 hCoV-19/England/ALDP-B73D06/2020\|EPI_ISL_664882\|20 | ···················· | ···················· | ···················· | ···················· | NNNNNNNNNNNNNNNNNNNN | NNNNN··············· | 1 |
| T | 3964 hCoV-19/England/ALDP-B73D42/2020\|EPI_ISL_664886\|20 | ···················· | ···················· | ···················· | ···················· | NNNNNNNNNNNNNNNNNNNN | NNNNNNNNNNNNNNNNNN·· | 1 |
| T | 3966 hCoV-19/England/ALDP-B73DAC/2020\|EPI_ISL_664888\|20 | ···················· | ···················· | ···················· | ···················· | NNNNNNNNNNNNNNNNNNNN | NNNNNNNNNNNNNNNNNNNN | 1 |
| T | 3967 hCoV-19/England/ALDP-B73DBB/2020\|EPI_ISL_664889\|20 | ···················· | ···················· | ···················· | ···················· | ···················· | NNNNN··············· | 1 |
| T | 3968 hCoV-19/England/ALDP-B73DCA/2020\|EPI_ISL_664890\|20 | ···················· | ···················· | ···················· | ···················· | NNNNNNNNNNNNNNNNNNNN | NNNNN··············· | 1 |
| T | 4170 hCoV-19/Australia/VIC16908/2020\|EPI_ISL_663530\|202 | ···················· | ···················· | ···················· | ···················· | NNNNNNNNNNNNNNNNNNNN | ···················· | 1 |
| T | 4171 hCoV-19/Australia/VIC16923/2020\|EPI_ISL_663531\|202 | ···················· | ···················· | ···················· | ···················· | NNNNNNNNNNNNNNNNNNNN | NNNNNNNNNNNNNNNNNNNN | 1 |
| T | 4175 hCoV-19/Australia/VIC16752/2020\|EPI_ISL_663535\|202 | ···················· | ···················· | ···················· | ···················· | NNNNNNNNNNNNNNNNNNNN | ···················· | 1 |
| T | 4177 hCoV-19/Australia/VIC16982/2020\|EPI_ISL_663537\|202 | ···················· | ···················· | ···················· | ···················· | NNNNNNNNNNNNNNNNNNNN | ···················· | 1 |
| T | 4179 hCoV-19/Australia/VIC17353/2020\|EPI_ISL_663539\|202 | ···················· | ···················· | ···················· | ···················· | NNNNNNNNNNNNNNNNNNNN | ···················· | 1 |
| T | 4181 hCoV-19/Australia/VIC17453/2020\|EPI_ISL_663541\|202 | ···················· | ···················· | ···················· | ···················· | NNNNNNNNNNNNNNNNNNNN | ···················· | 1 |
| T | 4184 hCoV-19/Australia/VIC16727/2020\|EPI_ISL_663553\|202 | ···················· | ···················· | ···················· | ···················· | NNNNNNNNNNNNNNNNNNNN | NNNNNNNNNNNNNNNNNNNN | 1 |
| T | 4209 hCoV-19/England/ALDP-B7398A/2020\|EPI_ISL_664863\|20 | ···················· | ···················· | ···················· | ···················· | NNNNNNNNNNNNNNNNNNNN | NNNNNNNNNNNNNNNNNNNN | 1 |
| T | 4211 hCoV-19/England/ALDP-B739C6/2020\|EPI_ISL_664865\|20 | ···················· | ···················· | ···················· | ···················· | NNNNNNNNNNNNNNNNNNNN | NNNNNNNNNNNNNNNNNNNN | 1 |
| T | 4212 hCoV-19/England/ALDP-B739D5/2020\|EPI_ISL_664866\|20 | ···················· | ···················· | ···················· | ···················· | NNN················· | NNNNN··············· | 1 |
| T | 4215 hCoV-19/England/ALDP-B73AB4/2020\|EPI_ISL_664869\|20 | ···················· | ···················· | ···················· | ···················· | NNNNNNNNNNNNNNNNNNNN | NNNNNNNNNNNNNNN····· | 1 |
| T | 4217 hCoV-19/England/ALDP-B73BDF/2020\|EPI_ISL_664871\|20 | ···················· | ···················· | ···················· | ···················· | NNNNNNNNNNNNNNNNNNNN | NNNNNNNNNNNNNNNNNNNN | 1 |
| T | 4220 hCoV-19/England/ALDP-B73C27/2020\|EPI_ISL_664874\|20 | ···················· | ···················· | ···················· | ···················· | NNNNNNNNNNNNNNNNNNNN | NNNNNNNNNNNNNNNNNNNN | 1 |
| T | 4391 hCoV-19/England/ALDP-B73841/2020\|EPI_ISL_664743\|20 | ···················· | ···················· | ···················· | ···················· | NNNNNNNNNNNNNNNNNNNN | NN·················· | 1 |
| T | 4393 hCoV-19/England/LOND-1250A0E/2020\|EPI_ISL_664745\|2 | ···················· | ···················· | ···················· | ···················· | NNNNNNNNNNNNNNNNNNNN | NNNNNNNNNNNNNNNNNNNN | 1 |
| T | 4397 hCoV-19/England/LOND-1250B1A/2020\|EPI_ISL_664755\|2 | ···················· | ···················· | ···················· | ···················· | NNNNNNNNNNNNNNNNNNNN | NNNNNNNNNNNNNNNNNNNN | 1 |
| T | 4445 hCoV-19/England/QEUH-B7C24A/2020\|EPI_ISL_668323\|20 | ···················· | NNNNNNNNNNNNNNNNNNNN | ···················· | ···················· | ···················· | ···················· | 1 |
| T | 4560 hCoV-19/Ireland/D-NVRL-75IRL77089/2020\|EPI_ISL_671 | ···················· | ···················· | ···················· | ···················· | NNNNNNNNNNNNNNNNNNNN | NNNNNNNNNNNNNNNNNNNN | 1 |
| T | 4563 hCoV-19/Ireland/LK-NVRL-75IRL32488/2020\|EPI_ISL_67 | ···················· | ···················· | ···················· | ···················· | NNNNNNNNNNNNNNNNNNNN | NNNNNNNNNNNNNNNNNNNN | 1 |
| T | 4678 hCoV-19/England/ALDP-B7396C/2020\|EPI_ISL_664741\|20 | ···················· | ···················· | ···················· | ···················· | NNNNNNNNNNNNNNNNNNNN | NNNNNNNNNNNNNNNNNNNN | 1 |
| T | 4686 hCoV-19/England/NOTT-11BD91/2020\|EPI_ISL_665900\|20 | ···················· | ···················· | ···················· | ···················· | NNNNNNNNNNNNNNNNNNNN | NNNNNNNNNNNNNNNNNNNN | 1 |
| T | 4689 hCoV-19/England/NOTT-11C60E/2020\|EPI_ISL_665903\|20 | ···················· | ···················· | ···················· | ···················· | NNNNNNNNNNNNNNNNNNNN | NNNNNNNNNNNNNNNNNNNN | 1 |
| T | 4719 hCoV-19/England/QEUH-B7E7EF/2020\|EPI_ISL_668142\|20 | ···················· | NNNNNNNNNNNNNNNNNNNN | ···················· | ···················· | ···················· | ···················· | 1 |
| T | 4748 hCoV-19/England/QEUH-B7C1C5/2020\|EPI_ISL_668197\|20 | ···················· | NNNNNNNNNNNNNNNNNNNN | ···················· | ···················· | ···················· | ···················· | 1 |
| T | 4749 hCoV-19/England/QEUH-B806DE/2020\|EPI_ISL_668198\|20 | ···················· | NNNNNNNNNNNNNNNNNNNN | ···················· | ···················· | ···················· | ···················· | 1 |
| T | 4750 hCoV-19/England/QEUH-B7C277/2020\|EPI_ISL_668199\|20 | ···················· | NNNNNNNNNNNNNNNNNNNN | ···················· | ···················· | ···················· | ···················· | 1 |
| T | 4761 hCoV-19/England/QEUH-B823F3/2020\|EPI_ISL_668216\|20 | ···················· | NNNNNNNNNNNNNNNNNNNN | ···················· | ···················· | ···················· | ···················· | 1 |
| T | 4837 hCoV-19/USA/CA-SCCPHD-UC197/2020\|EPI_ISL_671765\|20 | ···················· | ···················· | ···················· | ···················· | ··NNNNNNNNNNNNNNNNNN | NNNNNNNNNNNNNNNNNNNN | 1 |
| T | 4839 hCoV-19/Spain/AR-IBV-97009509/2020\|EPI_ISL_671797\| | ···················· | ···················· | ···················· | ···················· | NNNNNNNNNNNNNNNNNNNN | NNNNNNNNNNNNNNNNNNNN | 1 |
| T | 4840 hCoV-19/Spain/AR-IBV-97009511/2020\|EPI_ISL_671799\| | ···················· | ···················· | ···················· | ···················· | NNNNNNNNNNNNNNNNNNNN | NNNNNNNNNNNNNNNNNNNN | 1 |

T 4841 hCoV-19/Spain/CT-IBV-97009530/2020|EPI_ISL_671836| ···················· ···················· ···················· ···················· NNNNNNNNNNNNNNNNNNNN NNNNNNNNNNNNNNNNNNNN 1
T 4842 hCoV-19/Spain/CT-IBV-97009534/2020|EPI_ISL_671838| ···················· ···················· ···················· ···················· NNNNNNNNNNNNNNNNNNNN NNNNNNNNNNNNNNNNNNNN 1
T 4843 hCoV-19/Spain/CT-IBV-97009547/2020|EPI_ISL_671844| ···················· ···················· ···················· ···················· NNNNNNNNNNNNNNNNNNNN NNNNNNNNNNNNNNNNNNNN 1
T 4844 hCoV-19/Spain/CT-IBV-97009554/2020|EPI_ISL_671851| ···················· ···················· ···················· ···················· NNNNNNNNNNNNNNNNNNNN NNNNNNNNNNNNNNNNNNNN 1
T 4845 hCoV-19/Spain/CT-IBV-97009557/2020|EPI_ISL_671854| ···················· ···················· ···················· ···················· NNNNNNNNNNNNNNNNNNNN NNNNNNNNNNNNNNNNNNNN 1
T 4846 hCoV-19/Spain/CT-IBV-97009564/2020|EPI_ISL_671861| ···················· ···················· ···················· ···················· NNNNNNNNNNNNNNNNNNNN NNNNNNNNNNNNNNNNNNNN 1
T 4886 hCoV-19/France/NAQ-SC581/2020|EPI_ISL_663232|2020- ···················· ···················· ···················· ···················· NNNNNNNNNNNNNNNNNNNN ···················· 1
T 4895 hCoV-19/France/NAQ-SC598/2020|EPI_ISL_663242|2020- ···················· ········A··········· ···················· ···················· NNNNNNNNNNNNNNNNNNNN NNNNNNNNNNNNNNNNNNNN 1
T 4912 hCoV-19/France/PDL-SC613/2020|EPI_ISL_663259|2020- ···················· ···················· ···················· ···················· NNNNNNNNNNNNNNNNNNNN ···················· 1
T 4918 hCoV-19/France/PDL-SC622/2020|EPI_ISL_663265|2020- ···················· ···················· ···················· ···················· NNNN················ ···················· 1
T 4936 hCoV-19/England/EXET-1391CE/2020|EPI_ISL_664603|20 ···················· ···················· ···················· ···················· NNNNNNNNNNNNNNNNNNNN NNNNNNNNNNNNNNNNNNNN 1
T 4938 hCoV-19/England/NORW-F562A/2020|EPI_ISL_665887|202 ···················· ···················· ···················· ···················· NNNNNNNNNNNNN······· NNNNNNNNNNNNNNN····· 1
T 4939 hCoV-19/England/NORW-F5657/2020|EPI_ISL_665888|202 ···················· ···················· ···················· ···················· NNNNNNNNNNNNNNNNNNNN NNNNNNNNNNNNNNNNNNNN 1
T 4940 hCoV-19/England/NORW-F5675/2020|EPI_ISL_665889|202 ···················· ···················· ···················· ···················· NNNNNNNNNNNNNNNNNNNN NNNNNNNNNNNNNNNNNNNN 1
T 4941 hCoV-19/England/NORW-F5693/2020|EPI_ISL_665890|202 ···················· ···················· ···················· ···················· NNNNNNNNNNNNNNNNNNNN NNNNNNNNNNNNNNNNNNNN 1
T 4942 hCoV-19/England/NORW-F5718/2020|EPI_ISL_665891|202 ···················· ···················· ···················· ···················· NNNNNNNNNNNNNNNNNNNN NNNNNNNNNNNNNNNN···· 1
T 4943 hCoV-19/England/NORW-F5763/2020|EPI_ISL_665892|202 ···················· ···················· ···················· ···················· NNNNNNNNNNNNNNNNNNNN NNNNNNNNNNNNNNNNNNNN 1
T 4945 hCoV-19/England/NORW-F5824/2020|EPI_ISL_665894|202 ···················· ···················· ···················· ···················· ···················· NNNNNNNN·NN········· 1
T 4946 hCoV-19/England/NORW-F58BB/2020|EPI_ISL_665895|202 ···················· ···················· ···················· ···················· NNNNNNNNNNNNNNNNNNNN NNNNNNNNNNNNNNNNNNNN 1
T 4947 hCoV-19/England/NORW-F5CFB/2020|EPI_ISL_665896|202 ···················· ···················· ···················· ···················· NNNNNNNNNNNNNNNNNNNN NNNNNNNNNNNNNNNNNNNN 1
T 4948 hCoV-19/England/NORW-F745B/2020|EPI_ISL_665897|202 ···················· ···················· ···················· ···················· NNNNNNNNNNNNNNNNNNNN NNNNNNNNNNNNNNNNNNNN 1
T 4977 hCoV-19/England/PORT-2EA19E/2020|EPI_ISL_665946|20 ···················· ···················· ···················· ···················· NNNNNNNNNNNNNNNNNNNN NNNNNNNNNNNNNNNNNNNN 1
T 5485 hCoV-19/Wales/PHWC-48CDA7/2020|EPI_ISL_665779|2020 ···················· ···················· ···················· ···················· ···················· NNNNN··············· 1
T 5491 hCoV-19/England/NORW-F4F02/2020|EPI_ISL_665788|202 ···················· ···················· ···················· ···················· NNNNNNNNNNNNNNNNNNNN NNNNNNNNNNN········· 1
T 5495 hCoV-19/Wales/PHWC-48B133/2020|EPI_ISL_665793|2020 ···················· ···················· ···················· ···················· NNNN················ ···················· 1
T 5501 hCoV-19/England/LOND-1250C71/2020|EPI_ISL_665807|2 ···················· ·A·················· ···················· ···················· NNNNNNNNNNNNNNNNNNNN NNNNNNNNNNNNNNNNNNNN 1
T 5511 hCoV-19/England/LOND-1250DF6/2020|EPI_ISL_665817|2 ···················· ···················· ···················· ···················· NNNNNNNNNNNNNNNNN··· NNNNNNNNNNNNNNNNNNN· 1
T 5512 hCoV-19/England/LOND-1250E02/2020|EPI_ISL_665818|2 ···················· ···················· ···················· ···················· NNNNNNNNNNNN····N··· ···················· 1
T 5515 hCoV-19/England/LOND-1250E4E/2020|EPI_ISL_665821|2 ···················· ···················· ···················· ···················· ···················· NNNNNNNN············ 1
T 5520 hCoV-19/England/LOND-1250FB4/2020|EPI_ISL_665826|2 ···················· ···················· ···················· ···················· NNNNNNNNNNNN········ ···················· 1
T 5522 hCoV-19/England/LOND-1251029/2020|EPI_ISL_665828|2 ···················· ···················· ···················· ···················· ···················· NNNN················ 1
T 5530 hCoV-19/England/LOND-12511EA/2020|EPI_ISL_665836|2 ···················· ···················· ···················· ···················· NNNNNNNNNNNNNNNNNNNN ···················· 1
T 5533 hCoV-19/England/LOND-1251232/2020|EPI_ISL_665839|2 ···················· ···················· ···················· ···················· NNNNNNNNNNNNNNNNNNNN NNNNNNNNNNNNNNNNNNNN 1
T 5538 hCoV-19/England/LOND-1251320/2020|EPI_ISL_665844|2 ···················· ········A··········· ···················· ···················· ·····T·············· NNNNNNNNNNNNNNNNNNNN 1
T 5544 hCoV-19/England/MILK-A563F9/2020|EPI_ISL_665850|20 ···················· ···················· ···················· ···················· NNNNNNNNNNNNNNNNNNNN NNNNNNNNNNNNNNNNNNNN 1
T 5545 hCoV-19/England/MILK-A697BB/2020|EPI_ISL_665851|20 ···················· ···················· ···················· ···················· NNNNNNNNNNNNNNNNNNNN NNNNNNNNNNNNNNNNNNNN 1
T 5547 hCoV-19/England/MILK-A6985E/2020|EPI_ISL_665853|20 ···················· ·A·················· ···················· ···················· NNNNNNNNNNNNNNNNNNNN NNNNNNNNNNNNNNNNNNNN 1
T 5549 hCoV-19/England/MILK-A6992E/2020|EPI_ISL_665855|20 ···················· ···················· ···················· ···················· NNNNNNNNNNNNNNNNNNNN NNNNNNNNNNNNNNNNNNNN 1
T 5550 hCoV-19/England/MILK-A699B5/2020|EPI_ISL_665856|20 ···················· ···················· ···················· ···················· NNNNNNNNNNNNNNNNNNNN NNNNNNNNNNNNNNNNNNNN 1
T 5551 hCoV-19/England/MILK-A699F1/2020|EPI_ISL_665857|20 ···················· ···················· ···················· ···················· NNNNNNNNNNNNNNNNNNNN NNNNNNNNNNNNNNNNNNNN 1
T 5553 hCoV-19/England/MILK-A69A76/2020|EPI_ISL_665859|20 ···················· ···················· ···················· ···················· NNNNNNNNNNNNNNNNNNNN NNNNNNNNNNNNNNNNNNNN 1
T 5557 hCoV-19/England/MILK-A69B46/2020|EPI_ISL_665863|20 ···················· ···················· ···················· ···················· NNNNNNNNNNNNNNNNNNNN NNNNNNNNNNNNNNNNNNNN 1
T 5572 hCoV-19/England/NORW-F4ED8/2020|EPI_ISL_665878|202 ···················· ···················· ···················· ···················· NNNNNNNNNNNNNNNNNNNN NNN················· 1
T 5574 hCoV-19/England/NORW-F4FD5/2020|EPI_ISL_665880|202 ···················· ···················· ···················· ···················· NNNNNNNNNNNNNNNNNNNN NNNNNNNNNNNNNNNNNNNN 1
T 5580 hCoV-19/England/NORW-F5147/2020|EPI_ISL_665886|202 ···················· ···················· ···················· ···················· NNN··T·············· ···················· 1
T 5737 hCoV-19/Australia/VIC17477/2020|EPI_ISL_663900|202 ···················· ···················· ···················· ···················· NNNNNNNNNNNNNNNNNNNN ···················· 1
T 5803 hCoV-19/Australia/VIC17647/2020|EPI_ISL_663966|202 ···················· ···················· ···················· ···················· NNNNNNNNNNNNNNNNNNNN ···················· 1
T 5828 hCoV-19/Wales/PHWC-48B027/2020|EPI_ISL_666185|2020 ···················· ···················· ···················· ···················· NNNN················ ···················· 1
T 5834 hCoV-19/Wales/PHWC-48B0AF/2020|EPI_ISL_666191|2020 ···················· ···················· ···················· ···················· NNNNNNNNNNNNNNNNNNNN NNNNNNNNNNNNNNNNNNNN 1
T 5838 hCoV-19/Wales/PHWC-48B115/2020|EPI_ISL_666195|2020 ···················· ···················· ···················· ···················· NNNNNNNNNNNNNNNNNNNN NNNNNNNNNNNNNNNNNNNN 1
T 5954 hCoV-19/Australia/VIC17668/2020|EPI_ISL_663986|202 ···················· ···················· ···················· ···················· NNNNNNNNNNNNNNNNNNNN ···················· 1
T 5962 hCoV-19/Australia/VIC17680/2020|EPI_ISL_663994|202 ···················· ···················· ···················· ···················· NNNNNNNNNNNNNNNNNNNN NNNN················ 1
T 5964 hCoV-19/Australia/VIC17682/2020|EPI_ISL_663996|202 ···················· ···················· ···················· ···················· ···················· NNNNNNNNNNNNNNN····· 1
T 6000 hCoV-19/Wales/PHWC-48AC06/2020|EPI_ISL_666135|2020 ···················· ···················· ···················· ···················· NNNNNNNNNNNNNNNN···· NNNNNNNNNNN········· 1
T 6008 hCoV-19/Wales/PHWC-48ACAC/2020|EPI_ISL_666143|2020 ···················· ···················· ···················· ···················· NNNN················ ···················· 1
T 6018 hCoV-19/Wales/PHWC-48AD6D/2020|EPI_ISL_666153|2020 ···················· ···················· ···················· ···················· NNNNNNNNNNNNNNNNNNNN NNNNN··············· 1
T 6020 hCoV-19/Wales/PHWC-48AD9A/2020|EPI_ISL_666155|2020 ···················· ···················· ···················· ···················· NNNNNNNNNNNNNNNNNNNN ···················· 1
T 6036 hCoV-19/Wales/PHWC-48AEE2/2020|EPI_ISL_666171|2020 ···················· ···················· ···················· ···················· NNNNNNNNNNNNNNNNNNNN ···················· 1
T 6042 hCoV-19/Wales/PHWC-48AF58/2020|EPI_ISL_666177|2020 ···················· ···················· ···················· ···················· NNNN················ ···················· 1
T 6051 hCoV-19/USA/OR-OHSU-2908/2020|EPI_ISL_667407|2020- ···················· ···················· ···················· ···················· NNNNNNNNNNNNNNNNNNNN ···················· 1
T 6148 hCoV-19/New Zealand/20CV0588/2020|EPI_ISL_661262|2 ···················· ···················· ···················· ···················· NNNNNNNNNNNNNNNNNNNN NNNNNNNNNNNNNNNNNNNN 1
T 6149 hCoV-19/New Zealand/20CV0594/2020|EPI_ISL_661264|2 ···················· ·A·················· ···················· ···················· NNNNNNNNNNNNNNNNNNNN NNNNNNNNNNNNNNNNNNNN 1
T 6258 hCoV-19/Australia/VIC17247/2020|EPI_ISL_663812|202 ···················· ···················· ···················· ···················· NNNN················ ···················· 1
T 6259 hCoV-19/Australia/VIC17270/2020|EPI_ISL_663813|202 ···················· ···················· ···················· ···················· NNNNNNNNNNNNNNNNN··· NNNNNNNNN··········· 1
T 6260 hCoV-19/Australia/VIC17272/2020|EPI_ISL_663814|202 ···················· ···················· ···················· ···················· NNNNNNNNNNNNNNNNNNNN NNNNNNNNNNNNNNNNNNNN 1
T 6261 hCoV-19/Australia/VIC17273/2020|EPI_ISL_663815|202 ···················· ···················· ···················· ···················· NNNNNNNNNNNNNNNNN··· ···················· 1
T 6262 hCoV-19/Australia/VIC17274/2020|EPI_ISL_663816|202 ···················· ···················· ···················· ···················· NNNNNNNNNNNNNNNNNNNN ···················· 1
T 6263 hCoV-19/Australia/VIC17276/2020|EPI_ISL_663817|202 ···················· ···················· ···················· ···················· NNNNNNNNNNNNNNNNNNNN ···················· 1
T 6265 hCoV-19/Australia/VIC17283/2020|EPI_ISL_663819|202 ···················· ···················· ···················· ···················· NNNNNNNNNNNNNNNNNNNN ···················· 1
T 6266 hCoV-19/Australia/VIC17290/2020|EPI_ISL_663820|202 ···················· ···················· ···················· ···················· NNNNNNNNNNNNNNNNNNNN NNNNNNNNNNNNNNNNNNNN 1
T 6275 hCoV-19/Australia/VIC17304/2020|EPI_ISL_663829|202 ···················· ···················· ···················· ···················· NNNNNNNNNNNNNNNNNNNN NNNNNNNNNNNNNNNNNNNN 1
T 6286 hCoV-19/Australia/VIC17322/2020|EPI_ISL_663840|202 ···················· ···················· ···················· ···················· NNNNNNNNN··········· ···················· 1
T 6294 hCoV-19/Australia/VIC17335/2020|EPI_ISL_663848|202 ···················· ···················· ···················· ···················· NNNNNNNNNNNNNNNNNNNN NNNNNNNNNNNNNNNNNNNN 1
T 6297 hCoV-19/Australia/VIC17341/2020|EPI_ISL_663851|202 ···················· ···················· ···················· ···················· NNNNNNNNNNNNNNNNNNNN NNNNNNNNNNNNNNNNNNNN 1
T 6300 hCoV-19/Australia/VIC17354/2020|EPI_ISL_663854|202 ···················· ···················· ···················· ···················· ···················· NNNNNNNNNNNNNNN····· 1
T 6303 hCoV-19/Australia/VIC17363/2020|EPI_ISL_663857|202 ···················· ···················· ···················· ···················· NNNNNNNNNNNNNNNNNNNN NNNNNNNNNNNNNNN····· 1
T 6305 hCoV-19/Australia/VIC17365/2020|EPI_ISL_663859|202 ···················· ···················· ···················· ···················· NNNNNNNNNNNNNNNNNNNN ···················· 1
T 6307 hCoV-19/Australia/VIC17368/2020|EPI_ISL_663861|202 ···················· ···················· ···················· ···················· NNNNNNNNNNNNNNNNNNNN NNNNNNNNNNNNNNNNNNNN 1
T 6309 hCoV-19/Australia/VIC17374/2020|EPI_ISL_663863|202 ···················· ···················· ···················· ···················· NNNNNNNNNNNNNNNNNNNN ···················· 1
T 6312 hCoV-19/Australia/VIC17381/2020|EPI_ISL_663866|202 ···················· ···················· ···················· ···················· NNNNNNNNNNNNNNNNNNNN ··············NNNNNN 1

| | | | | | | | | | |
|---|---|---|---|---|---|---|---|---|---|
| T | 6321 | hCoV-19/Australia/VIC17408/2020\|EPI_ISL_663875\|202 | ···················· | ···················· | ···················· | ···················· | NNNNNNNNNNNNNNNNN··· | ···················· | 1 |
| T | 6324 | hCoV-19/Australia/VIC17415/2020\|EPI_ISL_663878\|202 | ···················· | ···················· | ···················· | ···················· | NNNNNNNNNNNNNNNNNNNN | ···················· | 1 |
| T | 6327 | hCoV-19/Australia/VIC17422/2020\|EPI_ISL_663881\|202 | ···················· | ···················· | ···················· | ···················· | NNNNNNNNNNNNNNNNNNNN | NNNNNNNNNNNNNNNNNNNN | 1 |
| T | 6328 | hCoV-19/Australia/VIC17428/2020\|EPI_ISL_663882\|202 | ···················· | ···················· | ···················· | ···················· | NNNNNNNNNNNNNNNNNNNN | NNNNNNNNNNNNNNNNNNNN | 1 |
| T | 6329 | hCoV-19/Australia/VIC17429/2020\|EPI_ISL_663883\|202 | ···················· | ···················· | ···················· | ···················· | NNNNN··············· | ···················· | 1 |
| T | 6331 | hCoV-19/Australia/VIC17433/2020\|EPI_ISL_663885\|202 | ···················· | ···················· | ···················· | ···················· | NNNNNNN············· | NNNNNNNNNNNNNNN····· | 1 |
| T | 6334 | hCoV-19/Australia/VIC17444/2020\|EPI_ISL_663888\|202 | ···················· | ···················· | ···················· | ···················· | NNNNNNNNNNNNNNNNNNNN | NNNNNNNNNNNNNNNNNNNN | 1 |
| T | 6336 | hCoV-19/Australia/VIC17446/2020\|EPI_ISL_663890\|202 | ···················· | ···················· | ···················· | ···················· | ················NNNN | ···················· | 1 |
| T | 6338 | hCoV-19/Australia/VIC17452/2020\|EPI_ISL_663892\|202 | ···················· | ···················· | ···················· | ···················· | ···················· | NNNNNNNNNNNNNNNNN··· | 1 |
| T | 6340 | hCoV-19/Australia/VIC17460/2020\|EPI_ISL_663894\|202 | ···················· | ···················· | ···················· | ···················· | NNNNNNN············· | ···················· | 1 |
| T | 6344 | hCoV-19/Australia/VIC17469/2020\|EPI_ISL_663898\|202 | ···················· | ···················· | ···················· | ···················· | NNNNNNNNNNNNNNNNNNNN | NNNNNNNNNNNNNNNNNNNN | 1 |
| T | 6348 | hCoV-19/Scotland/EDB9219/2020\|EPI_ISL_666062\|2020- | ···················· | ···················· | ···················· | ···················· | NNNNNNNNNNNNNNNNNNNN | NNNNNNNNNNNNNNNNNNNN | 1 |
| T | 6360 | hCoV-19/Scotland/EDB9317/2020\|EPI_ISL_666074\|2020- | ···················· | ···················· | ···················· | ···················· | NNNNNNNNNNNNNNNNNNNN | NNNNNNNNNNNNNNNNNNNN | 1 |
| T | 6361 | hCoV-19/Scotland/EDB9324/2020\|EPI_ISL_666075\|2020- | ···················· | ·········A·········· | ···················· | ···················· | NNNNNNNNNNNNNNNNNNNN | NNNNNNNNNNNNNNNNNNNN | 1 |
| T | 6362 | hCoV-19/Scotland/EDB9328/2020\|EPI_ISL_666076\|2020- | ···················· | ···················· | ···················· | ···················· | NNNNNNNNNNNNNNNNNNNN | NNNNNNNNNNNNNNNNNNNN | 1 |
| T | 6366 | hCoV-19/Scotland/EDB9349/2020\|EPI_ISL_666080\|2020- | ···················· | ···················· | ···················· | ···················· | NNNNNNNNNNNNNNNNNNNN | NNNNNNNNNNNNNNNNNNNN | 1 |
| T | 6451 | hCoV-19/USA/OR-OHSU-2767/2020\|EPI_ISL_667305\|2020- | ···················· | ···················· | ···················· | ···················· | ···················· | NNNNNNNNNNNNNNN····· | 1 |
| T | 6753 | hCoV-19/Australia/VIC17176/2020\|EPI_ISL_663779\|202 | ···················· | ···················· | ···················· | ···················· | NNNNNNNNNNNNNNNNNNNN | ···················· | 1 |
| T | 6754 | hCoV-19/Australia/VIC17179/2020\|EPI_ISL_663780\|202 | ···················· | ···················· | ···················· | ···················· | NNNNNNNNNNNNNNNNNNNN | ···················· | 1 |
| T | 6755 | hCoV-19/Australia/VIC17183/2020\|EPI_ISL_663781\|202 | ···················· | ···················· | ···················· | ···················· | NNNNNNNNNNNNNNNNNNNN | ···················· | 1 |
| T | 6757 | hCoV-19/Australia/VIC17187/2020\|EPI_ISL_663783\|202 | ···················· | ···················· | ···················· | ···················· | NNNNNNNNNNNNNNNNNNNN | NNNNNNNNNNNNNNNNNNNN | 1 |
| T | 7220 | hCoV-19/Australia/VIC16862/2020\|EPI_ISL_663611\|202 | ···················· | ···················· | ···················· | ···················· | ···················· | ····NNNN············ | 1 |
| T | 7222 | hCoV-19/Australia/VIC16868/2020\|EPI_ISL_663613\|202 | ···················· | ···················· | ···················· | ···················· | NNNNNNNNNNNNNNNNNNNN | ···················· | 1 |
| T | 7224 | hCoV-19/Australia/VIC16873/2020\|EPI_ISL_663615\|202 | ···················· | ···················· | ···················· | ···················· | NNNNNNNNNNNNNNNNNNNN | ···················· | 1 |
| T | 7228 | hCoV-19/Australia/VIC16880/2020\|EPI_ISL_663619\|202 | ···················· | ···················· | ···················· | ···················· | NNNNNNNNNNNNNNNNNNNN | NNNN··········NNNNNN | 1 |
| T | 7229 | hCoV-19/Australia/VIC16881/2020\|EPI_ISL_663620\|202 | ···················· | ···················· | ···················· | ···················· | NNNNNNNNNNNNNNNNNNNN | NNNNNNNNNNNNNNNNNNNN | 1 |
| T | 7233 | hCoV-19/Australia/VIC16947/2020\|EPI_ISL_663657\|202 | ···················· | ···················· | ···················· | ···················· | NNNNNNNNNNNNNNNNNNNN | ···················· | 1 |
| T | 7236 | hCoV-19/Australia/VIC16972/2020\|EPI_ISL_663667\|202 | ···················· | ···················· | ···················· | ···················· | NNNNNNNNNNNNNNNNNNNN | NNNNNNNNNNNNNNNNNNNN | 1 |
| T | 7237 | hCoV-19/Australia/VIC16975/2020\|EPI_ISL_663668\|202 | ···················· | ···················· | ···················· | ···················· | NNNNNNNNNNNNNNN····· | ···················· | 1 |
| T | 7240 | hCoV-19/Australia/VIC16984/2020\|EPI_ISL_663671\|202 | ···················· | ···················· | ···················· | ···················· | NNNNNNNNNNNN········ | NNNN················ | 1 |
| T | 7242 | hCoV-19/Australia/VIC16987/2020\|EPI_ISL_663673\|202 | ···················· | ···················· | ···················· | ···················· | NNNNNNNNNNNNNNNNNNNN | ···················· | 1 |
| T | 7258 | hCoV-19/Australia/VIC17023/2020\|EPI_ISL_663689\|202 | ···················· | ···················· | ···················· | ···················· | NNNN·····K···S·NNNNN | ···················· | 1 |
| T | 7271 | hCoV-19/England/ALDP-B74145/2020\|EPI_ISL_664902\|20 | ···················· | ···················· | ···················· | ···················· | NNNNNNNNNNNNNNNNNNNN | NNNNN··············· | 1 |
| T | 7276 | hCoV-19/England/ALDP-B741AF/2020\|EPI_ISL_664907\|20 | ···················· | ···················· | ···················· | ···················· | NNNNNNNNNNNNN······· | NNNNNNNNNNN········· | 1 |
| T | 7279 | hCoV-19/England/ALDP-B741DC/2020\|EPI_ISL_664910\|20 | ···················· | ···················· | ···················· | ···················· | NNNNNNNNNNNNNNNNNNNN | NNNNNNNNNNNNNNNNNNNN | 1 |
| T | 7288 | hCoV-19/England/ALDP-B74F69/2020\|EPI_ISL_664919\|20 | ···················· | ···················· | ···················· | ···················· | NNN················· | NNNNNNNNNNNNNNN····· | 1 |
| T | 7299 | hCoV-19/England/ALDP-B75A67/2020\|EPI_ISL_664930\|20 | ···················· | ···················· | ···················· | ···················· | NNNNNNNNNNNNNNNNNNNN | NNNNN··············· | 1 |
| T | 7302 | hCoV-19/England/ALDP-B75AB2/2020\|EPI_ISL_664933\|20 | ···················· | ···················· | ···················· | ···················· | NNNNNNNNNNNNNNNNNNNN | NNNNNNNNNNN········· | 1 |
| T | 7308 | hCoV-19/England/ALDP-B7733D/2020\|EPI_ISL_664939\|20 | ···················· | ···················· | ···················· | ···················· | NNNNNNNNNNN········· | NNNNN··············· | 1 |
| T | 7309 | hCoV-19/England/ALDP-B7734C/2020\|EPI_ISL_664940\|20 | ···················· | ···················· | ···················· | ···················· | NNNNNNNNNNNNNNNNNNNN | NNNNN··············· | 1 |
| T | 7480 | hCoV-19/Australia/VIC16828/2020\|EPI_ISL_663591\|202 | ···················· | ···················· | ···················· | ···················· | NNNNNNNNNNNNNNNNNNNN | ···················· | 1 |
| T | 7481 | hCoV-19/Australia/VIC16832/2020\|EPI_ISL_663592\|202 | ···················· | ···················· | ···················· | ···················· | NNNNNNN············· | ···················· | 1 |
| T | 7483 | hCoV-19/Australia/VIC16835/2020\|EPI_ISL_663594\|202 | ···················· | ···················· | ···················· | ···················· | NNNNNNNNNNNNNNNNN··· | ···················· | 1 |
| T | 7485 | hCoV-19/Australia/VIC16837/2020\|EPI_ISL_663596\|202 | ···················· | ···················· | ···················· | ···················· | NNNNNNNNNNNNNNNNNNNN | NNNNN·······NNN····· | 1 |
| T | 7489 | hCoV-19/Australia/VIC16882/2020\|EPI_ISL_663621\|202 | ···················· | ···················· | ···················· | ···················· | NNNNNNNNNNNNNNNNNNNN | NNNNNNNNNNNNNNNNNNNN | 1 |
| T | 7494 | hCoV-19/Australia/VIC16890/2020\|EPI_ISL_663626\|202 | ···················· | ···················· | ···················· | ···················· | NNNNNNNNNNNNNNNNNNNN | ··········NNNNN····· | 1 |
| T | 7495 | hCoV-19/Australia/VIC16893/2020\|EPI_ISL_663627\|202 | ···················· | ···················· | ···················· | ···················· | NNNNNNNNN··········· | ···················· | 1 |
| T | 7498 | hCoV-19/Australia/VIC16897/2020\|EPI_ISL_663630\|202 | ···················· | ···················· | ···················· | ···················· | NNNNNNNNNNNNNNNNNNNN | NNNNNNNNNNNNNNNNNNNN | 1 |
| T | 7504 | hCoV-19/Australia/VIC16912/2020\|EPI_ISL_663636\|202 | ···················· | ·····A·············· | ···················· | ···················· | NNNNNNNNNNNNNNNNNNNN | ····N··············· | 1 |
| T | 7506 | hCoV-19/Australia/VIC16914/2020\|EPI_ISL_663638\|202 | ···················· | ···················· | ···················· | ···················· | NNNNNNNNNNNNNNNNNNNN | NNNNNNNNNNNNNNNNNNNN | 1 |
| T | 7508 | hCoV-19/Australia/VIC16916/2020\|EPI_ISL_663640\|202 | ···················· | ···················· | ···················· | ···················· | NNNNNNNNNNNNNNNNNNNN | ···················· | 1 |
| T | 7513 | hCoV-19/Australia/VIC16932/2020\|EPI_ISL_663645\|202 | ···················· | ···················· | ···················· | ···················· | NNNNNNNNNNNNNNNNNN·· | ···················· | 1 |
| T | 7522 | hCoV-19/Australia/VIC16948/2020\|EPI_ISL_663658\|202 | ···················· | ···················· | ···················· | ···················· | NNNNNNNNNNNNNNNNNNNN | ···················· | 1 |
| T | 7524 | hCoV-19/Australia/VIC16957/2020\|EPI_ISL_663660\|202 | ···················· | ···················· | ···················· | ···················· | NNNNNNNN············ | ···················· | 1 |
| T | 7628 | hCoV-19/Sri Lanka/cov3563/2020\|EPI_ISL_668454\|2020 | ···················· | ···················· | ···················· | NNNNNNN············· | ···················· | ···················· | 1 |
| U | 196 | hCoV-19/Wales/PHWC-48E1D6/2020\|EPI_ISL_680821\|2020 | ···················· | ···················· | ···················· | ···················· | NNNNNNNNNNNNNNNNNNNN | NNNNNNNNNNNNNNNNNNNN | 1 |
| U | 197 | hCoV-19/Wales/PHWC-48E1E5/2020\|EPI_ISL_680822\|2020 | ···················· | ···················· | ···················· | ···················· | NNNNNNNNNNNNNNNNNNNN | NNNNNNNNNNNNNNNNNNNN | 1 |
| U | 204 | hCoV-19/Wales/PHWC-48E288/2020\|EPI_ISL_680832\|2020 | ···················· | ···················· | ···················· | ···················· | NNNNNNNNNNNNNNNNNNNN | NNNNNNNNNNNNNNNNNNNN | 1 |
| U | 205 | hCoV-19/Wales/PHWC-48E297/2020\|EPI_ISL_680833\|2020 | ···················· | ···················· | ···················· | ···················· | NNNNNNNNNNNNNNNNNNNN | NNNNNNNNNNNNNNNNNNNN | 1 |
| U | 207 | hCoV-19/Wales/PHWC-48E2B5/2020\|EPI_ISL_680835\|2020 | ···················· | ·····A·············· | ···················· | ···················· | NNNNNNNNNNNNNNNNNNNN | NNNNNNNNNNNNNNNNNNNN | 1 |
| U | 217 | hCoV-19/Wales/PHWC-48E385/2020\|EPI_ISL_680848\|2020 | ···················· | ···················· | ···················· | ···················· | NNNNNNNNNNNNNNNNNNNN | NNNNNNNNNNNNNNNNNNNN | 1 |
| U | 224 | hCoV-19/Wales/PHWC-48E419/2020\|EPI_ISL_680856\|2020 | ···················· | ···················· | ···················· | ···················· | NNNNNNNNNNNNNNNNNNNN | NNNNNNNNNNNNNNNNNNNN | 1 |
| U | 225 | hCoV-19/Wales/PHWC-48E428/2020\|EPI_ISL_680857\|2020 | ···················· | ···················· | ···················· | ···················· | NNNNNNNNNNNNNNNNNNNN | ···················· | 1 |
| U | 230 | hCoV-19/Wales/PHWC-48E491/2020\|EPI_ISL_680864\|2020 | ···················· | ···················· | ···················· | ···················· | NNNNNNNNNNNNNNNNNNNN | NNNNNNNNNNNNNNNNNNNN | 1 |
| U | 232 | hCoV-19/Wales/PHWC-48E4DD/2020\|EPI_ISL_680867\|2020 | ···················· | ···················· | ···················· | ···················· | NNNNNNNNNNNNNNNNNNNN | ···················· | 1 |
| U | 233 | hCoV-19/Wales/PHWC-48E4EC/2020\|EPI_ISL_680868\|2020 | ···················· | ···················· | ···················· | ···················· | ···················· | NNNNNNNNNNNNNNN····· | 1 |
| U | 241 | hCoV-19/Wales/PHWC-48E76B/2020\|EPI_ISL_680906\|2020 | ···················· | ···················· | ···················· | ···················· | NNNN················ | ···················· | 1 |
| U | 437 | hCoV-19/USA/CO-CDPHE-2004230027/2020\|EPI_ISL_67728 | ···················· | ···················· | ···················· | ···················· | NNNNNNNNNNNNNNNNNNNN | ···················· | 1 |
| U | 439 | hCoV-19/USA/CO-CDPHE-2004230144/2020\|EPI_ISL_67729 | ···················· | ···················· | ···················· | ···················· | NNNN················ | ···················· | 1 |
| U | 446 | hCoV-19/USA/CO-CDPHE-2007234527/2020\|EPI_ISL_67729 | ···················· | ···················· | ···················· | ···················· | NNNNNNNNNNNNNNNNNNNN | NNNN················ | 1 |
| U | 481 | hCoV-19/Wales/PHWC-48E516/2020\|EPI_ISL_680871\|2020 | ···················· | ···················· | ···················· | ···················· | N··················· | NNNNNNNNNNNNNNN····· | 1 |
| U | 482 | hCoV-19/Wales/PHWC-48E525/2020\|EPI_ISL_680872\|2020 | ···················· | ···················· | ···················· | ···················· | NNNNNNNNNNNNNNNNNNNN | ···················· | 1 |
| U | 486 | hCoV-19/Wales/PHWC-48E570/2020\|EPI_ISL_680877\|2020 | ···················· | ···················· | ···················· | ···················· | NNNNNNNNNNNNNNNNNNNN | NNNNNNNNNNNNNNNNNNNN | 1 |
| U | 488 | hCoV-19/Wales/PHWC-48E5CB/2020\|EPI_ISL_680881\|2020 | ···················· | ···················· | ···················· | ···················· | ···················· | NNNNNN·············· | 1 |
| U | 490 | hCoV-19/Wales/PHWC-48E5E9/2020\|EPI_ISL_680883\|2020 | ···················· | ···················· | ···················· | ···················· | NNNNNNNNNNNNNNNNNNNN | NNNNNNNNNNNNNNNNNNNN | 1 |
| U | 492 | hCoV-19/Wales/PHWC-48E613/2020\|EPI_ISL_680886\|2020 | ···················· | ···················· | ···················· | ···················· | ···················· | NNNNNNNNNNNNNNNNNNNN | 1 |
| U | 494 | hCoV-19/Wales/PHWC-48E631/2020\|EPI_ISL_680888\|2020 | ···················· | ···················· | ···················· | ···················· | NNNNNNNNNNNNNNNNNNNN | NNNNNNNNNNNNNNNNNNNN | 1 |
| U | 498 | hCoV-19/Wales/PHWC-48E6B9/2020\|EPI_ISL_680895\|2020 | ···················· | ···················· | ···················· | ···················· | NNNNNNNNNNNNNNNNNNNN | NNNNNNNNNNNNNNNNNNNN | 1 |
| U | 502 | hCoV-19/Wales/PHWC-48E6F5/2020\|EPI_ISL_680899\|2020 | ···················· | ···················· | ···················· | ···················· | NNNNNNNNNNNNNNNNNNNN | NNNNNNNNNNNNNNNNNNNN | 1 |
| U | 711 | hCoV-19/Wales/PHWC-48D9A2/2020\|EPI_ISL_680706\|2020 | ···················· | ···················· | ···················· | ···················· | NNNNNNNNNNNNNNNNNNNN | NNNNNNNNNNNNNNNNNNNN | 1 |

| | | | | | | | | | |
|---|---|---|---|---|---|---|---|---|---|
| U | 713 | hCoV-19/Wales/PHWC-48D9C0/2020\|EPI_ISL_680708\|2020 | ···················· | ···················· | ···················· | ···················· | NNNNNNNNNNNNNNNNNNNN | NNNNNNNNNNNNNNNNNNNN | 1 |
| U | 714 | hCoV-19/Wales/PHWC-48D9DF/2020\|EPI_ISL_680709\|2020 | ···················· | ···················· | ···················· | ···················· | NNNNNNNNNNNNNNNNNNNN | ···················· | 1 |
| U | 715 | hCoV-19/Wales/PHWC-48D9EE/2020\|EPI_ISL_680710\|2020 | ···················· | ···················· | ···················· | ···················· | NNNNNNNNNNNNNNNNNNNN | ···················· | 1 |
| U | 716 | hCoV-19/Wales/PHWC-48D9FD/2020\|EPI_ISL_680711\|2020 | ···················· | ···················· | ···················· | ···················· | NNNNNNNNNNNNNNNNNNNN | ·····NNNNNNNNNN····· | 1 |
| U | 717 | hCoV-19/Wales/PHWC-48DA09/2020\|EPI_ISL_680712\|2020 | ···················· | ···················· | ···················· | ···················· | NNNNNNNNNNNNNNNNNNNN | NNNNNNNNNNNNNNNNNNNN | 1 |
| U | 1011 | hCoV-19/Wales/PHWC-48DD97/2020\|EPI_ISL_680756\|2020 | ···················· | ···················· | ···················· | ···················· | NNNNNNNNNNNNNNNNNNNN | NNNNNNNNNNNNNNNNNNNN | 1 |
| U | 1200 | hCoV-19/Wales/PHWC-48D7F3/2020\|EPI_ISL_680682\|2020 | ···················· | ···················· | ···················· | ···················· | ··N················· | NNNNNNNNNNNNNNNNNNNN | 1 |
| U | 1481 | hCoV-19/Wales/PHWC-48D4A1/2020\|EPI_ISL_680631\|2020 | ···················· | ···················· | ···················· | ···················· | NNNNNNNNNNNNNN······ | ···················· | 1 |
| U | 1595 | hCoV-19/Saudi Arabia/KAUST-MAKKAH180/2020\|EPI_ISL_ | ···················· | ···················· | ···················· | ···················· | ······NNNNNNNNNNNNNN | NNNNNNNNNNNNNNNNNNNN | 1 |
| U | 1597 | hCoV-19/Saudi Arabia/KAUST-MAKKAH197/2020\|EPI_ISL_ | ···················· | ···················· | ···················· | ···················· | ······NNNNNNNNNNNNNN | NNNNNNNNNNNNNNNNNNNN | 1 |
| U | 1649 | hCoV-19/Scotland/EDB10208/2020\|EPI_ISL_680508\|2020 | ···················· | ···················· | ···················· | ···················· | NNNNNNNNNNNNNNNNNNNN | NNNNNNNNNNNNNNNNNNNN | 1 |
| U | 1651 | hCoV-19/Scotland/EDB10212/2020\|EPI_ISL_680510\|2020 | ···················· | ···················· | ···················· | ···················· | NNNNNNNNNNNNNNNNNNNN | NNNNNNNNNNNNNNNNNNNN | 1 |
| U | 1672 | hCoV-19/Scotland/EDB9446/2020\|EPI_ISL_680534\|2020- | ···················· | ···················· | ···················· | ···················· | NNNNNNNNNNNNNNNNNNNN | NNNNNNNNNNNNNNNNNNNN | 1 |
| U | 1677 | hCoV-19/Scotland/EDB9775/2020\|EPI_ISL_680540\|2020- | ···················· | ···················· | ···················· | ···················· | NNNNNNNNNNNNNNNNNNNN | NNNNNNNNNNNNNNNNNNNN | 1 |
| U | 1680 | hCoV-19/Scotland/EDB9818/2020\|EPI_ISL_680544\|2020- | ···················· | ···················· | ···················· | ···················· | NNNNNNNNNNNNNNNNNNNN | NNNNNNNNNNNNNNNNNNNN | 1 |
| U | 1698 | hCoV-19/Wales/PHWC-4862CC/2020\|EPI_ISL_680580\|2020 | ···················· | ···················· | ···················· | ···················· | NNNNNNNNNNNNNNNNNNNN | ···················· | 1 |
| U | 1699 | hCoV-19/Wales/PHWC-486350/2020\|EPI_ISL_680581\|2020 | ···················· | ···················· | ···················· | ···················· | ·NNNNNNNNNNN········ | ···················· | 1 |
| U | 1739 | hCoV-19/Cote dIvoire/BKE0349/2020\|EPI_ISL_681832\|2 | ···················· | ···················· | ···················· | ···················· | NNNNNNNNNNNNNNNNNNNN | NNNNNNNNNNNNNNNNNNNN | 1 |
| U | 1740 | hCoV-19/Cote dIvoire/BKE0356/2020\|EPI_ISL_681833\|2 | ···················· | ···················· | ···················· | ···················· | NNNNNNNNNNNNNNNNNNNN | NNNNNNNNNNNNNNNNNNNN | 1 |
| U | 1794 | hCoV-19/England/QEUH-BA1AAE/2020\|EPI_ISL_672851\|20 | ···················· | NNNNNNNNNNNNNNNNN··· | ···················· | ···················· | ···················· | ···················· | 1 |
| U | 1797 | hCoV-19/England/QEUH-BA11D6/2020\|EPI_ISL_672855\|20 | ···················· | NNNNNNNNNNNNNNNNNNNN | ···················· | ···················· | ···················· | ···················· | 1 |
| U | 1798 | hCoV-19/England/QEUH-BA264F/2020\|EPI_ISL_672856\|20 | ···················· | NNNNNNNNNNNNNNNNNNNN | ···················· | ···················· | ···················· | ···················· | 1 |
| U | 1818 | hCoV-19/England/QEUH-BA2C7A/2020\|EPI_ISL_672882\|20 | ···················· | NNNNNNNNNNNNNNNNNNNN | ···················· | ···················· | ···················· | ···················· | 1 |
| U | 1821 | hCoV-19/England/QEUH-BA2BE6/2020\|EPI_ISL_672888\|20 | ···················· | NNNNNNNNNNNNNNNNNNNN | ···················· | ···················· | ···················· | ···················· | 1 |
| U | 1899 | hCoV-19/USA/MI-UM-10036754004/2020\|EPI_ISL_682201\| | ···················· | ···················· | ···················· | ···················· | NNNNNNNNNNNNNNNNNNNN | NNNNNNNNNNNNNNNNNNNN | 1 |
| U | 1900 | hCoV-19/USA/MI-UM-10036746278/2020\|EPI_ISL_682202\| | ···················· | ·A·················· | ···················· | ···················· | NNNNNNNNNNNNNNNNNNNN | NNNNNNNNNNNNNNNNNNNN | 1 |
| U | 1901 | hCoV-19/USA/MI-UM-10036766200/2020\|EPI_ISL_682203\| | ···················· | ···················· | ···················· | ···················· | NNNNNNNNNNNNNNNNNNNN | NNNNNNNNNNNNNNNNNNNN | 1 |
| U | 1907 | hCoV-19/USA/MI-UM-10036767679/2020\|EPI_ISL_682209\| | ···················· | ···················· | ···················· | ···················· | ······NNNNNNNNNNNNNN | ···················· | 1 |
| U | 1908 | hCoV-19/USA/MI-UM-10036762912/2020\|EPI_ISL_682210\| | ···················· | ····A··············· | ···················· | ···················· | NNNNNNNNNNNNNNNNNNNN | ···················· | 1 |
| U | 1909 | hCoV-19/USA/MI-UM-10036758403/2020\|EPI_ISL_682211\| | ···················· | ······A············· | ···················· | ···················· | NNNNNNNNNNNNNNNNNNNN | ···················· | 1 |
| U | 1910 | hCoV-19/USA/MI-UM-10036764295/2020\|EPI_ISL_682212\| | ···················· | ···················· | ···················· | ···················· | NNNNNNNNNNNNNNNNNNNN | NNNNNNNNNNNNNNNNNNNN | 1 |
| U | 1911 | hCoV-19/USA/MI-UM-10036758386/2020\|EPI_ISL_682213\| | ···················· | ···················· | ···················· | ···················· | NNNNNNNNNNNNNNNNNNNN | NNNNNNNNNNNNNNNNNNNN | 1 |
| U | 1912 | hCoV-19/USA/MI-UM-10036753865/2020\|EPI_ISL_682215\| | ···················· | ···················· | ···················· | ···················· | NNNNNNNNNNNNNNNNNNNN | NNNNNNNNNNNNNNNNNNNN | 1 |
| U | 1913 | hCoV-19/USA/MI-UM-10036762218/2020\|EPI_ISL_682216\| | ···················· | ···················· | ···················· | ···················· | NNNNNNNNNNNNNNNNNNNN | ···················· | 1 |
| U | 1915 | hCoV-19/USA/MI-UM-10036764505/2020\|EPI_ISL_682218\| | ···················· | ···················· | ···················· | ···················· | NNNNNNNNNNNNNNNNNNNN | NNNNNNNNNNNNNNNNNNNN | 1 |
| U | 1919 | hCoV-19/USA/MI-UM-10036762637/2020\|EPI_ISL_682223\| | ···················· | ···················· | ···················· | ···················· | NNNNNNNNNNNNNNNNNNNN | ···················· | 1 |
| U | 1922 | hCoV-19/USA/MI-UM-10036765872/2020\|EPI_ISL_682226\| | ···················· | ···················· | ···················· | ···················· | ·······NNNNNNNNNNNNN | ···················· | 1 |
| U | 1926 | hCoV-19/USA/MI-UM-10036767393/2020\|EPI_ISL_682230\| | ···················· | ···················· | ···················· | ···················· | ··NNNNNNNNNNNNNNNNNN | ···················· | 1 |
| U | 1927 | hCoV-19/USA/MI-UM-10036767474/2020\|EPI_ISL_682232\| | ···················· | ·············A······ | ···················· | ···················· | NNNNNNNNNNNNNNNNNNNN | ···················· | 1 |
| U | 1928 | hCoV-19/USA/MI-UM-10036758395/2020\|EPI_ISL_682234\| | ···················· | ······A············· | ···················· | ···················· | NNNNNNNNNNNNNNNNNNNN | NNNNNNNNNNNNNNNNNNNN | 1 |
| U | 2004 | hCoV-19/England/204690308/2020\|EPI_ISL_683531\|2020 | ···················· | NNNNNNNNNNNNNNNNNNNN | ···················· | ···················· | ···················· | ···············R···· | 1 |
| U | 2047 | hCoV-19/Spain/AR-IBV-97009927/2020\|EPI_ISL_683595\| | ···················· | ···················· | ···················· | ···················· | NNNNNNNNNNNNNNNNNNNN | NNNNNNNNNNNNNNNNNNNN | 1 |
| U | 2048 | hCoV-19/Spain/CT-IBV-97010156/2020\|EPI_ISL_683598\| | ···················· | ···················· | ···················· | ···················· | NNNNNNNNNNNNNNNNNNNN | NNNNNNNNNNNNNNNNNNNN | 1 |
| U | 2049 | hCoV-19/England/QEUH-BA2A8F/2020\|EPI_ISL_672795\|20 | ···················· | NNNNNNNNNNNNNNNNNNNN | ···················· | ···················· | ···················· | ···················· | 1 |
| U | 2216 | hCoV-19/USA/WI-UW-2221/2020\|EPI_ISL_677624\|2020-09 | ···················· | ···················· | ···················· | ···················· | NNNNNNNNNNNNNNNNNNNN | NNNNNNNNNNNNNNNNNNNN | 1 |
| U | 2217 | hCoV-19/USA/IL-UW-2222/2020\|EPI_ISL_677625\|2020-09 | ···················· | ···················· | ···················· | ···················· | NNNNNNNNNNNNNNNNNNNN | NNNNNNNNNNNNNNNNNNNN | 1 |
| U | 2218 | hCoV-19/USA/WI-UW-2223/2020\|EPI_ISL_677626\|2020-10 | ···················· | ···················· | ···················· | ···················· | NNNNNNNNNNNNNNNNNNNN | NNNNNNNNNNNNNNNNNNNN | 1 |
| U | 2220 | hCoV-19/USA/WI-UW-2225/2020\|EPI_ISL_677628\|2020-10 | ···················· | ···················· | ···················· | ···················· | NNNNNNNNNNNNNNNNNNNN | NNNNNNNNNNNNNNNNNNNN | 1 |
| U | 2224 | hCoV-19/USA/WI-UW-2229/2020\|EPI_ISL_677632\|2020-11 | ···················· | ···················· | ···················· | ···················· | NNNNNNNNNNNNNNNNNNNN | NNNNNNNNNNNNNNNNNNNN | 1 |
| U | 2235 | hCoV-19/USA/CO-CDPHE-2009090322/2020\|EPI_ISL_67764 | ···················· | ·A·················· | ···················· | ···················· | ·NNNNNNNNNNNNNNNNNNN | NNNNNNNNNNNNNNNN·NNN | 1 |
| U | 2243 | hCoV-19/USA/WI-UW-2134/2020\|EPI_ISL_677662\|2020-10 | ···················· | ···················· | ···················· | ···················· | NNNNNNNNNNNNNNNNNNNN | NNNNNNNNNNNNNNNNNNNN | 1 |
| U | 2302 | hCoV-19/USA/MI-UM-10036738588/2020\|EPI_ISL_682133\| | ···················· | ···················· | ···················· | ···················· | NNNNNNNNNNNNNNNNNNNN | ···················· | 1 |
| U | 2308 | hCoV-19/England/QEUH-BA2B13/2020\|EPI_ISL_672759\|20 | ···················· | NNNNNNNNNNNNNNNNNNNN | ···················· | ···················· | ···················· | ···················· | 1 |
| U | 2317 | hCoV-19/England/QEUH-BA2C89/2020\|EPI_ISL_672773\|20 | ···················· | NNNNNNNNNNNNNNNNNNNN | ···················· | ···················· | ···················· | ···················· | 1 |
| U | 2371 | hCoV-19/USA/WI-UW-2145/2020\|EPI_ISL_677549\|2020-10 | ···················· | ···················· | ···················· | ···················· | NNNNNNNNNNNNNNNNNNNN | NNNNNNNNNNNNNNNNNNNN | 1 |
| U | 2372 | hCoV-19/USA/WI-UW-2147/2020\|EPI_ISL_677551\|2020-10 | ···················· | ···················· | ···················· | ···················· | NNNNNNNNNNNNNNNNNNNN | NNNNNNNNNNNNNNNNNNNN | 1 |
| U | 2373 | hCoV-19/USA/WI-UW-2148/2020\|EPI_ISL_677552\|2020-10 | ···················· | ···················· | ···················· | ···················· | NNNNNNNNNNNNNNNNNNNN | NNNNNNNNNNNNNNNNNNNN | 1 |
| U | 2374 | hCoV-19/USA/WI-UW-2151/2020\|EPI_ISL_677555\|2020-10 | ···················· | ···················· | ···················· | ···················· | NNNNNNNNNNNNNNNNNNNN | NNNNNNNNNNNNNNNNNNNN | 1 |
| U | 2375 | hCoV-19/USA/WI-UW-2155/2020\|EPI_ISL_677559\|2020-10 | ···················· | ···················· | ···················· | ···················· | NNNNNNNNNNNNNNNNNNNN | NNNNNNNNNNNNNNNNNNNN | 1 |
| U | 2376 | hCoV-19/USA/WI-UW-2157/2020\|EPI_ISL_677561\|2020-10 | ···················· | ···················· | ···················· | ···················· | NNNNNNNNNNNNNNNNNNNN | NNNNNNNNNNNNNNNNNNNN | 1 |
| U | 2377 | hCoV-19/USA/WI-UW-2161/2020\|EPI_ISL_677565\|2020-10 | ···················· | ···················· | ···················· | ···················· | NNNNNNNNNNNNNNNNNNNN | NNNNNNNNNNNNNNNNNNNN | 1 |
| U | 2378 | hCoV-19/USA/WI-UW-2162/2020\|EPI_ISL_677566\|2020-10 | ···················· | ···················· | ···················· | ···················· | NNNNNNNNNNNNNNNNNNNN | NNNNNNNNNNNNNNNNNNNN | 1 |
| U | 2379 | hCoV-19/USA/WI-UW-2167/2020\|EPI_ISL_677571\|2020-10 | ···················· | ···················· | ···················· | ···················· | NNNNNNNNNNNNNNNNNNNN | NNNNNNNNNNNNNNNNNNNN | 1 |
| U | 2380 | hCoV-19/USA/WI-UW-2172/2020\|EPI_ISL_677576\|2020-10 | ···················· | ···················· | ···················· | ···················· | NNNNNNNNNNNNNNNNNNNN | NNNNNNNNNNNNNNNNNNNN | 1 |
| U | 2381 | hCoV-19/USA/WI-UW-2173/2020\|EPI_ISL_677577\|2020-10 | ···················· | ···················· | ···················· | ···················· | NNNNNNNNNNNNNNNNNNNN | NNNNNNNNNNNNNNNNNNNN | 1 |
| U | 2382 | hCoV-19/USA/WI-UW-2183/2020\|EPI_ISL_677587\|2020-10 | ···················· | ···················· | ···················· | ···················· | NNNNNNNNNNNNNNNNNNNN | NNNNNNNNNNNNNNNNNNNN | 1 |
| U | 2386 | hCoV-19/USA/WI-UW-2200/2020\|EPI_ISL_677603\|2020-10 | ···················· | ···················· | ···················· | ···················· | NNNNNNNNNNNNNNNNNNNN | NNNNNNNNNNNNNNNNNNNN | 1 |
| U | 2387 | hCoV-19/USA/WI-UW-2201/2020\|EPI_ISL_677604\|2020-10 | ···················· | ···················· | ···················· | ···················· | NNNNNNNNNNNNNNNNNNNN | NNNNNNNNNNNNNNNNNNNN | 1 |
| U | 2400 | hCoV-19/USA/WI-UW-2215/2020\|EPI_ISL_677618\|2020-10 | ···················· | ···················· | ···················· | ···················· | NNNNNNNNNNNNNNNNNNNN | NNNNNNNNNNNNNNNNNNNN | 1 |
| U | 2436 | hCoV-19/England/ALDP-B73F4C/2020\|EPI_ISL_678917\|20 | ···················· | ···················· | ···················· | ···················· | NNNNNNNNNNNNNNNNNNN· | ···················· | 1 |
| U | 2480 | hCoV-19/USA/MI-UM-10036707893/2020\|EPI_ISL_682118\| | ···················· | ···················· | ···················· | ···················· | NNNNNNNNNNNNNNNNNNNN | ···················· | 1 |
| U | 2482 | hCoV-19/USA/MI-UM-10036719595/2020\|EPI_ISL_682134\| | ···················· | ···················· | ···················· | ···················· | NNNNNNNNNNNNNNNNNNNN | ···················· | 1 |
| U | 2483 | hCoV-19/USA/MI-UM-10036711761/2020\|EPI_ISL_682138\| | ···················· | ···················· | ···················· | ···················· | NNNNNNNNNNNNNNNNNNNN | NNNNNNNNNNNNNNNNNNNN | 1 |
| U | 2493 | hCoV-19/USA/MI-UM-10036705785/2020\|EPI_ISL_682151\| | ···················· | ···················· | ···················· | ···················· | NNNNNNNNNNNNNNNNNNNN | ···················· | 1 |
| U | 2494 | hCoV-19/USA/MI-UM-10036721022/2020\|EPI_ISL_682152\| | ···················· | ···················· | ···················· | ···················· | NNNNNNNNNNNNNNNNNNNN | ···················· | 1 |
| U | 2495 | hCoV-19/USA/MI-UM-10036710728/2020\|EPI_ISL_682153\| | ···················· | ···················· | ···················· | ···················· | NNNNNNNNNNNNNNNNNNNN | ···················· | 1 |
| U | 2496 | hCoV-19/USA/MI-UM-10036721151/2020\|EPI_ISL_682157\| | ···················· | ···················· | ···················· | ···················· | NNNNNNNNNNNNNNNNNNNN | ···················· | 1 |
| U | 2507 | hCoV-19/USA/MI-UM-10036744194/2020\|EPI_ISL_682168\| | ···················· | ···················· | ···················· | ···················· | NNNNNNNNNNNNNNNNNNNN | NNNNNNNNNNNNNNNNNNNN | 1 |
| U | 2508 | hCoV-19/USA/MI-UM-10036737345/2020\|EPI_ISL_682169\| | ···················· | ·A·················· | ···················· | ···················· | NNNNNNNNNNNNNNNNNNNN | NNNNNNNNNNNNNNNNNNNN | 1 |
| U | 2530 | hCoV-19/USA/MI-UM-10036755231/2020\|EPI_ISL_682193\| | ···················· | ···················· | ···················· | ···················· | NNNNNNNNNNNNNNNNNNNN | ···················· | 1 |

| | | | | | | | | | |
|---|---|---|---|---|---|---|---|---|---|
| U | 2535 | hCoV-19/USA/MI-UM-10036756355/2020\|EPI_ISL_682198\| | ···················· | ···················· | ···················· | ···················· | NNNNNNNNNNNNNNNNNNNN | ···················· | 1 |
| U | 2536 | hCoV-19/USA/MI-UM-10036787296/2020\|EPI_ISL_682199\| | ···················· | ·A·················· | ···················· | ···················· | NNNNNNNNNNNNNNNNNNNN | ···················· | 1 |
| U | 2539 | hCoV-19/France/ARA-99504/2020\|EPI_ISL_683402\|2020- | ···················· | ···················· | ···················· | ···················· | NNNNNNNNNNNNNNNNNNNN | NNNNNNNNNNNNNNNNNNNN | 1 |
| U | 2679 | hCoV-19/USA/WI-UW-2096/2020\|EPI_ISL_677501\|2020-10 | ···················· | ···················· | ···················· | ···················· | NNNNNNNNNNNNNNNNNNNN | NNNNNNNNNNNNNNNNNNNN | 1 |
| U | 2680 | hCoV-19/USA/WI-UW-2097/2020\|EPI_ISL_677502\|2020-10 | ···················· | ···················· | ···················· | ···················· | NNNNNNNNNNNNNNNNNNNN | NNNNNNNNNNNNNNNNNNNN | 1 |
| U | 2681 | hCoV-19/USA/WI-UW-2098/2020\|EPI_ISL_677503\|2020-10 | ···················· | ···················· | ···················· | ···················· | NNNNNNNNNNNNNNNNNNNN | NNNNNNNNNNNNNNNNNNNN | 1 |
| U | 2682 | hCoV-19/USA/WI-UW-2099/2020\|EPI_ISL_677504\|2020-10 | ···················· | ···················· | ···················· | ···················· | NNNNNNNNNNNNNNNNNNNN | NNNNNNNNNNNNNNNNNNNN | 1 |
| U | 2683 | hCoV-19/USA/WI-UW-2100/2020\|EPI_ISL_677505\|2020-11 | ···················· | ···················· | ···················· | ···················· | NNNNNNNNNNNNNNNNNNNN | NNNNNNNNNNNNNNNNNNNN | 1 |
| U | 2685 | hCoV-19/USA/WI-UW-2102/2020\|EPI_ISL_677507\|2020-11 | ···················· | ···················· | ···················· | ···················· | NNNNNNNNNNNNNNNNNNNN | NNNNNNNNNNNNNNNNNNNN | 1 |
| U | 2687 | hCoV-19/USA/WI-UW-2104/2020\|EPI_ISL_677509\|2020-09 | ···················· | ···················· | ···················· | ···················· | NNNNNNNNNNNNNNNNNNNN | NNNNNNNNNNNNNNNNNNNN | 1 |
| U | 2688 | hCoV-19/USA/WI-UW-2105/2020\|EPI_ISL_677510\|2020-09 | ···················· | ···················· | ···················· | ···················· | NNNNNNNNNNNNNNNNNNNN | NNNNNNNNNNNNNNNNNNNN | 1 |
| U | 2689 | hCoV-19/USA/WI-UW-2106/2020\|EPI_ISL_677511\|2020-09 | ···················· | ···················· | ···················· | ···················· | NNNNNNNNNNNNNNNNNNNN | NNNNNNNNNNNNNNNNNNNN | 1 |
| U | 2690 | hCoV-19/USA/WI-UW-2107/2020\|EPI_ISL_677512\|2020-09 | ···················· | ···················· | ···················· | ···················· | NNNNNNNNNNNNNNNNNNNN | NNNNNNNNNNNNNNNNNNNN | 1 |
| U | 2691 | hCoV-19/USA/WI-UW-2108/2020\|EPI_ISL_677513\|2020-10 | ···················· | ···················· | ···················· | ···················· | NNNNNNNNNNNNNNNNNNNN | NNNNNNNNNNNNNNNNNNNN | 1 |
| U | 2692 | hCoV-19/USA/WI-UW-2109/2020\|EPI_ISL_677514\|2020-09 | ···················· | ···················· | ···················· | ···················· | NNNNNNNNNNNNNNNNNNNN | NNNNNNNNNNNNNNNNNNNN | 1 |
| U | 2693 | hCoV-19/USA/WI-UW-2110/2020\|EPI_ISL_677515\|2020-09 | ···················· | ···················· | ···················· | ···················· | NNNNNNNNNNNNNNNNNNNN | NNNNNNNNNNNNNNNNNNNN | 1 |
| U | 2694 | hCoV-19/USA/WI-UW-2111/2020\|EPI_ISL_677516\|2020-09 | ···················· | ···················· | ···················· | ···················· | NNNNNNNNNNNNNNNNNNNN | NNNNNNNNNNNNNNNNNNNN | 1 |
| U | 2695 | hCoV-19/USA/IL-UW-2112/2020\|EPI_ISL_677517\|2020-09 | ···················· | ···················· | ···················· | ···················· | NNNNNNNNNNNNNNNNNNNN | NNNNNNNNNNNNNNNNNNNN | 1 |
| U | 2697 | hCoV-19/USA/NE-UW-2116/2020\|EPI_ISL_677521\|2020-10 | ···················· | ···················· | ···················· | ···················· | NNNNNNNNNNNNNNNNNNNN | NNNNNNNNNNNNNNNNNNNN | 1 |
| U | 2699 | hCoV-19/USA/WI-UW-2118/2020\|EPI_ISL_677523\|2020-10 | ···················· | ···················· | ···················· | ···················· | NNNNNNNNNNNNNNNNNNNN | NNNNNNNNNNNNNNNNNNNN | 1 |
| U | 2700 | hCoV-19/USA/IL-UW-2119/2020\|EPI_ISL_677524\|2020-10 | ···················· | ···················· | ···················· | ···················· | NNNNNNNNNNNNNNNNNNNN | NNNNNNNNNNNNNNNNNNNN | 1 |
| U | 2701 | hCoV-19/USA/WI-UW-2120/2020\|EPI_ISL_677525\|2020-09 | ···················· | ···················· | ···················· | ···················· | NNNNNNNNNNNNNNNNNNNN | NNNNNNNNNNNNNNNNNNNN | 1 |
| U | 2702 | hCoV-19/USA/WI-UW-2121/2020\|EPI_ISL_677526\|2020-10 | ···················· | ···················· | ···················· | ···················· | NNNNNNNNNNNNNNNNNNNN | NNNNNNNNNNNNNNNNNNNN | 1 |
| U | 2704 | hCoV-19/USA/IL-UW-2123/2020\|EPI_ISL_677528\|2020-09 | ···················· | ···················· | ···················· | ···················· | NNNNNNNNNNNNNNNNNNNN | NNNNNNNNNNNNNNNNNNNN | 1 |
| U | 2705 | hCoV-19/USA/WI-UW-2124/2020\|EPI_ISL_677529\|2020-09 | ···················· | ···················· | ···················· | ···················· | NNNNNNNNNNNNNNNNNNNN | NNNNNNNNNNNNNNNNNNNN | 1 |
| U | 2706 | hCoV-19/USA/WI-UW-2126/2020\|EPI_ISL_677531\|2020-09 | ···················· | ···················· | ···················· | ···················· | NNNNNNNNNNNNNNNNNNNN | NNNNNNNNNNNNNNNNNNNN | 1 |
| U | 2707 | hCoV-19/USA/WI-UW-2127/2020\|EPI_ISL_677532\|2020-09 | ···················· | ···················· | ···················· | ···················· | NNNNNNNNNNNNNNNNNNNN | NNNNNNNNNNNNNNNNNNNN | 1 |
| U | 2708 | hCoV-19/USA/WI-UW-2128/2020\|EPI_ISL_677533\|2020-09 | ···················· | ···················· | ···················· | ···················· | NNNNNNNNNNNNNNNNNNNN | NNNNNNNNNNNNNNNNNNNN | 1 |
| U | 2709 | hCoV-19/USA/WI-UW-2129/2020\|EPI_ISL_677534\|2020-10 | ···················· | ···················· | ···················· | ···················· | NNNNNNNNNNNNNNNNNNNN | NNNNNNNNNNNNNNNNNNNN | 1 |
| U | 2711 | hCoV-19/USA/WI-UW-2131/2020\|EPI_ISL_677536\|2020-09 | ···················· | ···················· | ···················· | ···················· | NNNNNNNNNNNNNNNNNNNN | NNNNNNNNNNNNNNNNNNNN | 1 |
| U | 2712 | hCoV-19/USA/WI-UW-2135/2020\|EPI_ISL_677539\|2020-10 | ···················· | ···················· | ···················· | ···················· | NNNNNNNNNNNNNNNNNNNN | NNNNNNNNNNNNNNNNNNNN | 1 |
| U | 2713 | hCoV-19/USA/WI-UW-2140/2020\|EPI_ISL_677544\|2020-10 | ···················· | ···················· | ···················· | ···················· | NNNNNNNNNNNNNNNNNNNN | NNNNNNNNNNNNNNNNNNNN | 1 |
| U | 2714 | hCoV-19/USA/WI-UW-2141/2020\|EPI_ISL_677545\|2020-10 | ···················· | ···················· | ···················· | ···················· | NNNNNNNNNNNNNNNNNNNN | NNNNNNNNNNNNNNNNNNNN | 1 |
| U | 2715 | hCoV-19/USA/WI-UW-2142/2020\|EPI_ISL_677546\|2020-10 | ···················· | ···················· | ···················· | ···················· | NNNNNNNNNNNNNNNNNNNN | NNNNNNNNNNNNNNNNNNNN | 1 |
| U | 2717 | hCoV-19/USA/WI-UW-2189/2020\|EPI_ISL_677592\|2020-10 | ···················· | ···················· | ···················· | ···················· | NNNNNNNNNNNNNNNNNNNN | NNNNNNNNNNNNNNNNNNNN | 1 |
| U | 2719 | hCoV-19/USA/WI-UW-2191/2020\|EPI_ISL_677594\|2020-10 | ···················· | ···················· | ···················· | ···················· | NNNNNNNNNNNNNNNNNNNN | NNNNNNNNNNNNNNNNNNNN | 1 |
| U | 2724 | hCoV-19/USA/WI-UW-2196/2020\|EPI_ISL_677599\|2020-10 | ···················· | ···················· | ···················· | ···················· | NNNNNNNNNNNNNNNNNNNN | NNNNNNNNNNNNNNNNNNNN | 1 |
| U | 2784 | hCoV-19/USA/MI-UM-10036684811/2020\|EPI_ISL_682086\| | ···················· | ···················· | ···················· | ···················· | NNNNNNNNNNNNNNNNNNNN | ···················· | 1 |
| U | 2785 | hCoV-19/USA/MI-UM-10036682652/2020\|EPI_ISL_682087\| | ···················· | ···················· | ···················· | ···················· | ··NNNNNNNNNNNNNNNNNN | ···················· | 1 |
| U | 2797 | hCoV-19/mink/Denmark/mDK-570/2020\|EPI_ISL_683306\|2 | ···················· | ·A·················· | ···················· | ···················· | NNNNNNNNNNNNNNNNNNNN | NNNNNNNNNNNNNNNNNNNN | 1 |
| U | 2913 | hCoV-19/USA/WI-UW-2016/2020\|EPI_ISL_677423\|2020-10 | ···················· | ···················· | ···················· | ···················· | NNNNNNNNNNNNNNNNNNNN | NNNNNNNNNNNNNNNNNNNN | 1 |
| U | 2914 | hCoV-19/USA/WI-UW-2017/2020\|EPI_ISL_677424\|2020-10 | ···················· | ···················· | ···················· | ···················· | NNNNNNNNNNNNNNNNNNNN | NNNNNNNNNNNNNNNNNNNN | 1 |
| U | 2915 | hCoV-19/USA/IL-UW-2018/2020\|EPI_ISL_677425\|2020-10 | ···················· | ···················· | ···················· | ···················· | NNNNNNNNNNNNNNNNNNNN | NNNNNNNNNNNNNNNNNNNN | 1 |
| U | 2916 | hCoV-19/USA/WI-UW-2019/2020\|EPI_ISL_677426\|2020-09 | ···················· | ···················· | ···················· | ···················· | NNNNNNNNNNNNNNNNNNNN | NNNNNNNNNNNNNNNNNNNN | 1 |
| U | 2917 | hCoV-19/USA/WI-UW-2020/2020\|EPI_ISL_677427\|2020-09 | ···················· | ···················· | ···················· | ···················· | NNNNNNNNNNNNNNNNNNNN | NNNNNNNNNNNNNNNNNNNN | 1 |
| U | 2918 | hCoV-19/USA/WI-UW-2021/2020\|EPI_ISL_677428\|2020-09 | ···················· | ···················· | ···················· | ···················· | NNNNNNNNNNNNNNNNNNNN | NNNNNNNNNNNNNNNNNNNN | 1 |
| U | 2919 | hCoV-19/USA/WI-UW-2022/2020\|EPI_ISL_677429\|2020-10 | ···················· | ···················· | ···················· | ···················· | NNNNNNNNNNNNNNNNNNNN | NNNNNNNNNNNNNNNNNNNN | 1 |
| U | 2920 | hCoV-19/USA/WI-UW-2023/2020\|EPI_ISL_677430\|2020-10 | ···················· | ···················· | ···················· | ···················· | NNNNNNNNNNNNNNNNNNNN | NNNNNNNNNNNNNNNNNNNN | 1 |
| U | 2921 | hCoV-19/USA/IL-UW-2024/2020\|EPI_ISL_677431\|2020-09 | ···················· | ···················· | ···················· | ···················· | NNNNNNNNNNNNNNNNNNNN | NNNNNNNNNNNNNNNNNNNN | 1 |
| U | 2922 | hCoV-19/USA/WI-UW-2025/2020\|EPI_ISL_677432\|2020-09 | ···················· | ···················· | ···················· | ···················· | NNNNNNNNNNNNNNNNNNNN | NNNNNNNNNNNNNNNNNNNN | 1 |
| U | 2923 | hCoV-19/USA/WI-UW-2026/2020\|EPI_ISL_677433\|2020-10 | ···················· | ···················· | ···················· | ···················· | NNNNNNNNNNNNNNNNNNNN | NNNNNNNNNNNNNNNNNNNN | 1 |
| U | 2924 | hCoV-19/USA/WI-UW-2027/2020\|EPI_ISL_677434\|2020-09 | ···················· | ···················· | ···················· | ···················· | NNNNNNNNNNNNNNNNNNNN | NNNNNNNNNNNNNNNNNNNN | 1 |
| U | 2925 | hCoV-19/USA/WI-UW-2028/2020\|EPI_ISL_677435\|2020-09 | ···················· | ···················· | ···················· | ···················· | NNNNNNNNNNNNNNNNNNNN | NNNNNNNNNNNNNNNNNNNN | 1 |
| U | 2926 | hCoV-19/USA/WI-UW-2029/2020\|EPI_ISL_677436\|2020-09 | ···················· | ···················· | ···················· | ···················· | NNNNNNNNNNNNNNNNNNNN | NNNNNNNNNNNNNNNNNNNN | 1 |
| U | 2927 | hCoV-19/USA/IL-UW-2031/2020\|EPI_ISL_677437\|2020-09 | ···················· | ···················· | ···················· | ···················· | NNNNNNNNNNNNNNNNNNNN | NNNNNNNNNNNNNNNNNNNN | 1 |
| U | 2928 | hCoV-19/USA/WI-UW-2032/2020\|EPI_ISL_677438\|2020-10 | ···················· | ···················· | ···················· | ···················· | NNNNNNNNNNNNNNNNNNNN | NNNNNNNNNNNNNNNNNNNN | 1 |
| U | 2929 | hCoV-19/USA/IL-UW-2033/2020\|EPI_ISL_677439\|2020-09 | ···················· | ···················· | ···················· | ···················· | NNNNNNNNNNNNNNNNNNNN | NNNNNNNNNNNNNNNNNNNN | 1 |
| U | 2931 | hCoV-19/USA/WI-UW-2035/2020\|EPI_ISL_677441\|2020-10 | ···················· | ···················· | ···················· | ···················· | NNNNNNNNNNNNNNNNNNNN | NNNNNNNNNNNNNNNNNNNN | 1 |
| U | 2932 | hCoV-19/USA/WI-UW-2036/2020\|EPI_ISL_677442\|2020-09 | ···················· | ···················· | ···················· | ···················· | NNNNNNNNNNNNNNNNNNNN | NNNNNNNNNNNNNNNNNNNN | 1 |
| U | 2933 | hCoV-19/USA/WI-UW-2037/2020\|EPI_ISL_677443\|2020-09 | ···················· | ···················· | ···················· | ···················· | NNNNNNNNNNNNNNNNNNNN | NNNNNNNNNNNNNNNNNNNN | 1 |
| U | 2934 | hCoV-19/USA/WI-UW-2038/2020\|EPI_ISL_677444\|2020-09 | ···················· | ···················· | ···················· | ···················· | NNNNNNNNNNNNNNNNNNNN | NNNNNNNNNNNNNNNNNNNN | 1 |
| U | 2935 | hCoV-19/USA/WI-UW-2040/2020\|EPI_ISL_677446\|2020-10 | ···················· | ···················· | ···················· | ···················· | NNNNNNNNNNNNNNNNNNNN | NNNNNNNNNNNNNNNNNNNN | 1 |
| U | 2936 | hCoV-19/USA/WI-UW-2041/2020\|EPI_ISL_677447\|2020-10 | ···················· | ···················· | ···················· | ···················· | NNNNNNNNNNNNNNNNNNNN | NNNNNNNNNNNNNNNNNNNN | 1 |
| U | 2937 | hCoV-19/USA/WI-UW-2042/2020\|EPI_ISL_677448\|2020-09 | ···················· | ···················· | ···················· | ···················· | NNNNNNNNNNNNNNNNNNNN | NNNNNNNNNNNNNNNNNNNN | 1 |
| U | 2938 | hCoV-19/USA/WI-UW-2043/2020\|EPI_ISL_677449\|2020-10 | ···················· | ···················· | ···················· | ···················· | NNNNNNNNNNNNNNNNNNNN | NNNNNNNNNNNNNNNNNNNN | 1 |
| U | 2939 | hCoV-19/USA/IL-UW-2045/2020\|EPI_ISL_677450\|2020-09 | ···················· | ···················· | ···················· | ···················· | NNNNNNNNNNNNNNNNNNNN | NNNNNNNNNNNNNNNNNNNN | 1 |
| U | 2940 | hCoV-19/USA/IL-UW-2046/2020\|EPI_ISL_677451\|2020-10 | ···················· | ·N·················· | ···················· | ···················· | NNNNNNNNNNNNNNNNNNNN | NNNNNNNNNNNNNNNNNNNN | 1 |
| U | 2941 | hCoV-19/USA/WI-UW-2048/2020\|EPI_ISL_677453\|2020-09 | ···················· | ···················· | ···················· | ···················· | NNNNNNNNNNNNNNNNNNNN | NNNNNNNNNNNNNNNNNNNN | 1 |
| U | 2942 | hCoV-19/USA/WI-UW-2049/2020\|EPI_ISL_677454\|2020-10 | ···················· | ···················· | ···················· | ···················· | NNNNNNNNNNNNNNNNNNNN | NNNNNNNNNNNNNNNNNNNN | 1 |
| U | 2945 | hCoV-19/USA/WI-UW-2052/2020\|EPI_ISL_677457\|2020-10 | ···················· | ···················· | ···················· | ···················· | NNNNNNNNNNNNNNNNNNNN | NNNNNNNNNNNNNNNNNNNN | 1 |
| U | 2950 | hCoV-19/USA/WI-UW-2057/2020\|EPI_ISL_677462\|2020-09 | ···················· | ···················· | ···················· | ···················· | NNNNNNNNNNNNNNNNNNNN | NNNNNNNNNNNNNNNNNNNN | 1 |
| U | 2952 | hCoV-19/USA/WI-UW-2059/2020\|EPI_ISL_677464\|2020-09 | ···················· | ···················· | ···················· | ···················· | NNNNNNNNNNNNNNNNNNNN | NNNNNNNNNNNNNNNNNNNN | 1 |
| U | 2953 | hCoV-19/USA/WI-UW-2060/2020\|EPI_ISL_677465\|2020-10 | ···················· | ···················· | ···················· | ···················· | NNNNNNNNNNNNNNNNNNNN | NNNNNNNNNNNNNNNNNNNN | 1 |
| U | 2954 | hCoV-19/USA/WI-UW-2061/2020\|EPI_ISL_677466\|2020-09 | ···················· | ···················· | ···················· | ···················· | NNNNNNNNNNNNNNNNNNNN | NNNNNNNNNNNNNNNNNNNN | 1 |
| U | 3187 | hCoV-19/USA/WI-UW-1911/2020\|EPI_ISL_677320\|2020-10 | ···················· | ···················· | ···················· | ···················· | NNNNNNNNNNNNNNNNNNNN | NNNNNNNNNNNNNNNNNNNN | 1 |
| U | 3189 | hCoV-19/USA/WI-UW-1913/2020\|EPI_ISL_677322\|2020-10 | ···················· | ···················· | ···················· | ···················· | NNNNNNNNNNNNNNNNNNNN | NNNNNNNNNNNNNNNNNNNN | 1 |
| U | 3190 | hCoV-19/USA/WI-UW-1914/2020\|EPI_ISL_677323\|2020-10 | ···················· | ···················· | ···················· | ···················· | NNNNNNNNNNNNNNNNNNNN | NNNNNNNNNNNNNNNNNNNN | 1 |
| U | 3191 | hCoV-19/USA/WI-UW-1915/2020\|EPI_ISL_677324\|2020-10 | ···················· | ···················· | ···················· | ···················· | NNNNNNNNNNNNNNNNNNNN | NNNNNNNNNNNNNNNNNNNN | 1 |
| U | 3192 | hCoV-19/USA/WI-UW-1916/2020\|EPI_ISL_677325\|2020-10 | ···················· | ···················· | ···················· | ···················· | NNNNNNNNNNNNNNNNNNNN | NNNNNNNNNNNNNNNNNNNN | 1 |
| U | 3193 | hCoV-19/USA/WI-UW-1917/2020\|EPI_ISL_677326\|2020-10 | ···················· | ···················· | ···················· | ···················· | NNNNNNNNNNNNNNNNNNNN | NNNNNNNNNNNNNNNNNNNN | 1 |

| | | | | | | | | | |
|---|---|---|---|---|---|---|---|---|---|
| U | 3194 | hCoV-19/USA/WI-UW-1918/2020\|EPI_ISL_677327\|2020-10 | ···················· | ···················· | ···················· | ···················· | NNNNNNNNNNNNNNNNNNNN | NNNNNNNNNNNNNNNNNNNN | 1 |
| U | 3195 | hCoV-19/USA/WI-UW-1919/2020\|EPI_ISL_677328\|2020-10 | ···················· | ···················· | ···················· | ···················· | NNNNNNNNNNNNNNNNNNNN | NNNNNNNNNNNNNNNNNNNN | 1 |
| U | 3196 | hCoV-19/USA/WI-UW-1920/2020\|EPI_ISL_677329\|2020-10 | ···················· | ···················· | ···················· | ···················· | NNNNNNNNNNNNNNNNNNNN | NNNNNNNNNNNNNNNNNNNN | 1 |
| U | 3197 | hCoV-19/USA/WI-UW-1921/2020\|EPI_ISL_677330\|2020-10 | ···················· | ···················· | ···················· | ···················· | NNNNNNNNNNNNNNNNNNNN | NNNNNNNNNNNNNNNNNNNN | 1 |
| U | 3198 | hCoV-19/USA/WI-UW-1922/2020\|EPI_ISL_677331\|2020-10 | ···················· | ···················· | ···················· | ···················· | NNNNNNNNNNNNNNNNNNNN | NNNNNNNNNNNNNNNNNNNN | 1 |
| U | 3199 | hCoV-19/USA/WI-UW-1923/2020\|EPI_ISL_677332\|2020-10 | ···················· | ···················· | ···················· | ···················· | NNNNNNNNNNNNNNNNNNNN | NNNNNNNNNNNNNNNNNNNN | 1 |
| U | 3201 | hCoV-19/USA/WI-UW-1925/2020\|EPI_ISL_677334\|2020-10 | ···················· | ···················· | ···················· | ···················· | NNNNNNNNNNNNNNNNNNNN | NNNNNNNNNNNNNNNNNNNN | 1 |
| U | 3206 | hCoV-19/USA/WI-UW-1931/2020\|EPI_ISL_677340\|2020-10 | ···················· | ···················· | ···················· | ···················· | NNNNNNNNNNNNNNNNNNNN | NNNNNNNNNNNNNNNNNNNN | 1 |
| U | 3212 | hCoV-19/USA/IL-UW-1937/2020\|EPI_ISL_677346\|2020-09 | ···················· | ···················· | ···················· | ···················· | NNNNNNNNNNNNNNNNNNNN | NNNNNNNNNNNNNNNNNNNN | 1 |
| U | 3219 | hCoV-19/USA/WI-UW-2069/2020\|EPI_ISL_677474\|2020-09 | ···················· | ···················· | ···················· | ···················· | NNNNNNNNNNNNNNNNNNNN | NNNNNNNNNNNNNNNNNNNN | 1 |
| U | 3221 | hCoV-19/USA/IL-UW-2071/2020\|EPI_ISL_677476\|2020-10 | ···················· | ···················· | ···················· | ···················· | NNNNNNNNNNNNNNNNNNNN | NNNNNNNNNNNNNNNNNNNN | 1 |
| U | 3226 | hCoV-19/USA/IL-UW-2076/2020\|EPI_ISL_677481\|2020-10 | ···················· | ·N·················· | ···················· | ···················· | NNNNNNNNNNNNNNNNNNNN | NNNNNNNNNNNNNNNNNNNN | 1 |
| U | 3227 | hCoV-19/USA/WI-UW-2077/2020\|EPI_ISL_677482\|2020-10 | ···················· | ···················· | ···················· | ···················· | NNNNNNNNNNNNNNNNNNNN | NNNNNNNNNNNNNNNNNNNN | 1 |
| U | 3229 | hCoV-19/USA/IL-UW-2079/2020\|EPI_ISL_677484\|2020-10 | ···················· | ···················· | ···················· | ···················· | NNNNNNNNNNNNNNNNNNNN | NNNNNNNNNNNNNNNNNNNN | 1 |
| U | 3230 | hCoV-19/USA/IL-UW-2080/2020\|EPI_ISL_677485\|2020-10 | ···················· | ···················· | ···················· | ···················· | NNNNNNNNNNNNNNNNNNNN | NNNNNNNNNNNNNNNNNNNN | 1 |
| U | 3231 | hCoV-19/USA/WI-UW-2081/2020\|EPI_ISL_677486\|2020-10 | ···················· | ···················· | ···················· | ···················· | NNNNNNNNNNNNNNNNNNNN | NNNNNNNNNNNNNNNNNNNN | 1 |
| U | 3234 | hCoV-19/USA/WI-UW-2084/2020\|EPI_ISL_677489\|2020-10 | ···················· | ···················· | ···················· | ···················· | NNNNNNNNNNNNNNNNNNNN | NNNNNNNNNNNNNNNNNNNN | 1 |
| U | 3235 | hCoV-19/USA/IL-UW-2085/2020\|EPI_ISL_677490\|2020-09 | ···················· | ···················· | ···················· | ···················· | NNNNNNNNNNNNNNNNNNNN | NNNNNNNNNNNNNNNNNNNN | 1 |
| U | 3241 | hCoV-19/USA/WI-UW-2091/2020\|EPI_ISL_677496\|2020-09 | ···················· | ···················· | ···················· | ···················· | NNNNNNNNNNNNNNNNNNNN | NNNNNNNNNNNNNNNNNNNN | 1 |
| U | 3242 | hCoV-19/USA/WI-UW-2092/2020\|EPI_ISL_677497\|2020-09 | ···················· | ···················· | ···················· | ···················· | NNNNNNNNNNNNNNNNNNNN | NNNNNNNNNNNNNNNNNNNN | 1 |
| U | 3291 | hCoV-19/Wales/PHWC-48EAEA/2020\|EPI_ISL_680961\|2020 | ···················· | ···················· | ···················· | ···················· | NNNNNNNNNNNNNNNNNNNN | NNNNNN·············· | 1 |
| U | 3292 | hCoV-19/Wales/PHWC-48EAF9/2020\|EPI_ISL_680962\|2020 | ···················· | ···················· | ···················· | ···················· | NNNNNNNNNNNNNNNNNNNN | ····NN·············· | 1 |
| U | 3293 | hCoV-19/Wales/PHWC-48EB14/2020\|EPI_ISL_680964\|2020 | ···················· | ···················· | ···················· | ···················· | NNNNNNNNNNNNNNNNNNNN | NNNNNNNNNNNNNNNNNNNN | 1 |
| U | 3295 | hCoV-19/Wales/PHWC-48EB41/2020\|EPI_ISL_680967\|2020 | ···················· | ···················· | ···················· | ···················· | ··NNNNNNNNNN·NNNNNNN | ···················· | 1 |
| U | 3419 | hCoV-19/USA/CO-CDPHE-2008170097/2020\|EPI_ISL_67730 | ···················· | ···················· | ···················· | ···················· | NNNNNNNNNNNNNNNNNNNN | NNNNNNNNNNNNNNNNNNNN | 1 |
| U | 3421 | hCoV-19/USA/CO-CDPHE-2008170563/2020\|EPI_ISL_67730 | ···················· | ···················· | ···················· | ···················· | NNNNNNNNNNNNNNNNNNNN | NNNNNNNNNNNNNNNNNNNN | 1 |
| U | 3425 | hCoV-19/USA/CO-CDPHE-2009151294/2020\|EPI_ISL_67731 | ···················· | ···················· | ···················· | ···················· | NNNNNNNNNNNNNNNNNNNN | ··············N····· | 1 |
| U | 3433 | hCoV-19/USA/WI-UW-1948/2020\|EPI_ISL_677356\|2020-09 | ···················· | ···················· | ···················· | ···················· | NNNNNNNNNNNNNNNNNNNN | NNNNNNNNNNNNNNNNNNNN | 1 |
| U | 3434 | hCoV-19/USA/WI-UW-1949/2020\|EPI_ISL_677357\|2020-09 | ···················· | ···················· | ···················· | ···················· | NNNNNNNNNNNNNNNNNNNN | NNNNNNNNNNNNNNNNNNNN | 1 |
| U | 3436 | hCoV-19/USA/WI-UW-1951/2020\|EPI_ISL_677359\|2020-09 | ···················· | ···················· | ···················· | ···················· | NNNNNNNNNNNNNNNNNNNN | NNNNNNNNNNNNNNNNNNNN | 1 |
| U | 3438 | hCoV-19/USA/WI-UW-1953/2020\|EPI_ISL_677361\|2020-09 | ···················· | ···················· | ···················· | ···················· | NNNNNNNNNNNNNNNNNNNN | NNNNNNNNNNNNNNNNNNNN | 1 |
| U | 3441 | hCoV-19/USA/IL-UW-1956/2020\|EPI_ISL_677364\|2020-09 | ···················· | ···················· | ···················· | ···················· | NNNNNNNNNNNNNNNNNNNN | NNNNNNNNNNNNNNNNNNNN | 1 |
| U | 3445 | hCoV-19/USA/IL-UW-1960/2020\|EPI_ISL_677368\|2020-09 | ···················· | ···················· | ···················· | ···················· | NNNNNNNNNNNNNNNNNNNN | NNNNNNNNNNNNNNNNNNNN | 1 |
| U | 3446 | hCoV-19/USA/IL-UW-1961/2020\|EPI_ISL_677369\|2020-09 | ···················· | ···················· | ···················· | ···················· | NNNNNNNNNNNNNNNNNNNN | NNNNNNNNNNNNNNNNNNNN | 1 |
| U | 3450 | hCoV-19/USA/WI-UW-1965/2020\|EPI_ISL_677373\|2020-10 | ···················· | ···················· | ···················· | ···················· | NNNNNNNNNNNNNNNNNNNN | NNNNNNNNNNNNNNNNNNNN | 1 |
| U | 3454 | hCoV-19/USA/WI-UW-1970/2020\|EPI_ISL_677378\|2020-09 | ···················· | ···················· | ···················· | ···················· | NNNNNNNNNNNNNNNNNNNN | NNNNNNNNNNNNNNNNNNNN | 1 |
| U | 3455 | hCoV-19/USA/WI-UW-1971/2020\|EPI_ISL_677379\|2020-09 | ···················· | ···················· | ···················· | ···················· | NNNNNNNNNNNNNNNNNNNN | NNNNNNNNNNNNNNNNNNNN | 1 |
| U | 3456 | hCoV-19/USA/WI-UW-1972/2020\|EPI_ISL_677380\|2020-09 | ···················· | ·············N······ | ···················· | ···················· | NNNNNNNNNNNNNNNNNNNN | NNNNNNNNNNNNNNNNNNNN | 1 |
| U | 3457 | hCoV-19/USA/WI-UW-1974/2020\|EPI_ISL_677382\|2020-09 | ···················· | ···················· | ···················· | ···················· | NNNNNNNNNNNNNNNNNNNN | NNNNNNNNNNNNNNNNNNNN | 1 |
| U | 3458 | hCoV-19/USA/WI-UW-1975/2020\|EPI_ISL_677383\|2020-09 | ···················· | ···················· | ···················· | ···················· | NNNNNNNNNNNNNNNNNNNN | NNNNNNNNNNNNNNNNNNNN | 1 |
| U | 3459 | hCoV-19/USA/WI-UW-1976/2020\|EPI_ISL_677384\|2020-09 | ···················· | ···················· | ···················· | ···················· | NNNNNNNNNNNNNNNNNNNN | NNNNNNNNNNNNNNNNNNNN | 1 |
| U | 3460 | hCoV-19/USA/WI-UW-1979/2020\|EPI_ISL_677387\|2020-09 | ···················· | ···················· | ···················· | ···················· | NNNNNNNNNNNNNNNNNNNN | NNNNNNNNNNNNNNNNNNNN | 1 |
| U | 3461 | hCoV-19/USA/WI-UW-1980/2020\|EPI_ISL_677388\|2020-09 | ···················· | ···················· | ···················· | ···················· | NNNNNNNNNNNNNNNNNNNN | NNNNNNNNNNNNNNNNNNNN | 1 |
| U | 3462 | hCoV-19/USA/IL-UW-1981/2020\|EPI_ISL_677389\|2020-09 | ···················· | ···················· | ···················· | ···················· | NNNNNNNNNNNNNNNNNNNN | NNNNNNNNNNNNNNNNNNNN | 1 |
| U | 3463 | hCoV-19/USA/WI-UW-1982/2020\|EPI_ISL_677390\|2020-09 | ···················· | ···················· | ···················· | ···················· | NNNNNNNNNNNNNNNNNNNN | NNNNNNNNNNNNNNNNNNNN | 1 |
| U | 3464 | hCoV-19/USA/IL-UW-1983/2020\|EPI_ISL_677391\|2020-09 | ···················· | ···················· | ···················· | ···················· | NNNNNNNNNNNNNNNNNNNN | NNNNNNNNNNNNNNNNNNNN | 1 |
| U | 3466 | hCoV-19/USA/WI-UW-1985/2020\|EPI_ISL_677393\|2020-09 | ···················· | ···················· | ···················· | ···················· | NNNNNNNNNNNNNNNNNNNN | NNNNNNNNNNNNNNNNNNNN | 1 |
| U | 3467 | hCoV-19/USA/WI-UW-1986/2020\|EPI_ISL_677394\|2020-09 | ···················· | ···················· | ·N·················· | ···················· | NNNNNNNNNNNNNNNNNNNN | NNNNNNNNNNNNNNNNNNNN | 1 |
| U | 3468 | hCoV-19/USA/WI-UW-1987/2020\|EPI_ISL_677395\|2020-09 | ···················· | ···················· | ···················· | ···················· | NNNNNNNNNNNNNNNNNNNN | NNNNNNNNNNNNNNNNNNNN | 1 |
| U | 3469 | hCoV-19/USA/WI-UW-1989/2020\|EPI_ISL_677397\|2020-09 | ···················· | ···················· | ···················· | ···················· | NNNNNNNNNNNNNNNNNNNN | NNNNNNNNNNNNNNNNNNNN | 1 |
| U | 3470 | hCoV-19/USA/WI-UW-1990/2020\|EPI_ISL_677398\|2020-09 | ···················· | ···················· | ···················· | ···················· | NNNNNNNNNNNNNNNNNNNN | NNNNNNNNNNNNNNNNNNNN | 1 |
| U | 3471 | hCoV-19/USA/WI-UW-1991/2020\|EPI_ISL_677399\|2020-09 | ···················· | ···················· | ···················· | ···················· | NNNNNNNNNNNNNNNNNNNN | NNNNNNNNNNNNNNNNNNNN | 1 |
| U | 3493 | hCoV-19/South Africa/KRISP-EC-K005341/2020\|EPI_ISL | ···················· | ···················· | ···················· | ···················· | ··NNNNNNNNNNNNNNNNNN | NNNNNNNNNNNNNNNNNNNN | 1 |
| U | 3494 | hCoV-19/South Africa/KRISP-EC-K005347/2020\|EPI_ISL | ···················· | ···················· | ···················· | ···················· | NNNNNNNNNNNNNNNNNNNN | NNNNNNNNNNNNNNNNNNNN | 1 |
| U | 3536 | hCoV-19/Wales/PHWC-48E7A7/2020\|EPI_ISL_680910\|2020 | ···················· | ···················· | ···················· | ···················· | NNNNNNNNNNNNNNNNNNNN | NNNNNNNNNNNNNNNNNNNN | 1 |
| U | 3537 | hCoV-19/Wales/PHWC-48E7B6/2020\|EPI_ISL_680911\|2020 | ···················· | ···················· | ···················· | ···················· | NNNNNNNNNNNNNNNNNNNN | ···················· | 1 |
| U | 3538 | hCoV-19/Wales/PHWC-48E7C5/2020\|EPI_ISL_680912\|2020 | ···················· | ···················· | ···················· | ···················· | NNNNNNNNNNNNNNNNNNNN | ···················· | 1 |
| U | 3547 | hCoV-19/Wales/PHWC-48E8A4/2020\|EPI_ISL_680925\|2020 | ···················· | ···················· | ···················· | ···················· | ···················· | NNNN················ | 1 |
| U | 3548 | hCoV-19/Wales/PHWC-48E8B3/2020\|EPI_ISL_680926\|2020 | ···················· | ···················· | ···················· | ···················· | NNNNNNNNNNNNNNNNNNNN | NNNNNNNNNNNNNNNNNNNN | 1 |
| U | 3550 | hCoV-19/Wales/PHWC-48E8D1/2020\|EPI_ISL_680928\|2020 | ···················· | ···················· | ···················· | ···················· | NNNNNNNNNNNNNNNNNNNN | NNNNNNNNNNNNNNNNNNNN | 1 |
| U | 3552 | hCoV-19/Wales/PHWC-48E8FF/2020\|EPI_ISL_680930\|2020 | ···················· | ···················· | ···················· | ···················· | NNNNNNNNNNNNNNNNNNNN | NNNNNNNNNNNNNNNNNNNN | 1 |
| U | 3555 | hCoV-19/Wales/PHWC-48E929/2020\|EPI_ISL_680933\|2020 | ···················· | ···················· | ···················· | ···················· | NNNNNNNNNNNNNNNNNNNN | NNNNNNNNNNNNNNNNNNNN | 1 |
| U | 3558 | hCoV-19/Wales/PHWC-48E983/2020\|EPI_ISL_680939\|2020 | ···················· | ···················· | ···················· | ···················· | NNNN················ | ···················· | 1 |
| U | 3560 | hCoV-19/Wales/PHWC-48E9A1/2020\|EPI_ISL_680941\|2020 | ···················· | ···················· | ···················· | ···················· | NNNNNNNNNNNNNNNNNNNN | NNNNNNNNNNNNNNNNNNNN | 1 |
| U | 3561 | hCoV-19/Wales/PHWC-48E9B0/2020\|EPI_ISL_680942\|2020 | ···················· | ···················· | ···················· | ···················· | NNN················· | NNNNN··············· | 1 |
| U | 3562 | hCoV-19/Wales/PHWC-48E9ED/2020\|EPI_ISL_680945\|2020 | ···················· | ···················· | ···················· | ···················· | NNNNNNNNNNNNNNNNNNNN | NNNNNNNNNNNNNNNNNNNN | 1 |
| U | 3563 | hCoV-19/Wales/PHWC-48E9FC/2020\|EPI_ISL_680946\|2020 | ···················· | ···················· | ···················· | ···················· | NNNNNNNNNNNNNNNNNNNN | NNNNNNNNNNNNNNNNNNNN | 1 |
| U | 3567 | hCoV-19/Wales/PHWC-48EA35/2020\|EPI_ISL_680950\|2020 | ···················· | ···················· | ···················· | ···················· | NNNNNNNNNNNNNNNNNNNN | NNNNNNNNNNNNNNNNNNNN | 1 |
| U | 3571 | hCoV-19/Wales/PHWC-48EA80/2020\|EPI_ISL_680955\|2020 | ···················· | ···················· | ···················· | ···················· | NNNNNNNNNNNNNNNNNNNN | ···················· | 1 |
| U | 3575 | hCoV-19/Wales/PHWC-48EACC/2020\|EPI_ISL_680959\|2020 | ···················· | ···················· | ···················· | ···················· | NNNNNNNNNNNNNNNNNNNN | NNNNNNNNNNNNNNNNNNNN | 1 |
| U | 3727 | hCoV-19/USA/WI-UW-1617/2020\|EPI_ISL_681320\|2020-09 | ···················· | ···················· | ···················· | ···················· | NNNNNNNNNNNNNNNNNNNN | NNNNNNNNNNNNNNNNNNNN | 1 |
| U | 4111 | hCoV-19/Wales/PHWC-48FCC5/2020\|EPI_ISL_681214\|2020 | ···················· | ···················· | ···················· | ···················· | NNNNNNNNNNNNNNNNNNNN | NNNNN······NNNN····· | 1 |
| U | 4112 | hCoV-19/Wales/PHWC-48FCD4/2020\|EPI_ISL_681215\|2020 | ···················· | ···················· | ···················· | ···················· | NNNNNNNNNNNNNNNNNNNN | ····NNNNNNNNNNN·NNNN | 1 |
| U | 4113 | hCoV-19/Wales/PHWC-48FCE3/2020\|EPI_ISL_681216\|2020 | ···················· | ···················· | ···················· | ···················· | NNNNNNNNNNNNNNNNNNNN | NNNNNNNNNNNNNNNNNNNN | 1 |
| U | 4123 | hCoV-19/Wales/PHWC-48FDC2/2020\|EPI_ISL_681229\|2020 | ···················· | ···················· | ···················· | ···················· | NNNNNNNNNNNNNNNNNNNN | NNNNNNNNNNNNNNNNNNNN | 1 |
| U | 4125 | hCoV-19/Wales/PHWC-48FDE0/2020\|EPI_ISL_681231\|2020 | ···················· | ···················· | ···················· | ···················· | NNNNNNNNNNNNNNNNNNNN | ···················· | 1 |
| U | 4128 | hCoV-19/Wales/PHWC-48FE29/2020\|EPI_ISL_681235\|2020 | ···················· | ···················· | ···················· | ···················· | ···················· | NNNNNNNNNNNNNNN····· | 1 |
| U | 4131 | hCoV-19/Wales/PHWC-48FE56/2020\|EPI_ISL_681238\|2020 | ···················· | ···················· | ···················· | ···················· | NNNNNNNNNNNNNNNNNNNN | NNNNNNNNNNNNNNNNNNNN | 1 |
| U | 4132 | hCoV-19/Wales/PHWC-48FE65/2020\|EPI_ISL_681239\|2020 | ···················· | ···················· | ···················· | ···················· | NNNNNNNNNNNNNNNNNNNN | NNNNNNNNNNNNNNNNNNNN | 1 |
| U | 4133 | hCoV-19/Wales/PHWC-48FE92/2020\|EPI_ISL_681242\|2020 | ···················· | ···················· | ···················· | ···················· | NNNNNNNNNNNNNNNNNNNN | NNNNNNNNNNNNNNNNNNNN | 1 |

| | | | | | | | | | |
|---|---|---|---|---|---|---|---|---|---|
| U | 4137 | hCoV-19/Wales/PHWC-48FEDE/2020\|EPI_ISL_681246\|2020 | ···················· | ···················· | ···················· | ···················· | NNNNNNNNNNNNNNNNNNNN | ···················· | 1 |
| U | 4139 | hCoV-19/Wales/PHWC-48FF17/2020\|EPI_ISL_681249\|2020 | ···················· | ···················· | ···················· | ···················· | ···················· | NNNNNNNNNNNNNNN····· | 1 |
| U | 4147 | hCoV-19/Wales/PHWC-48FFBD/2020\|EPI_ISL_681259\|2020 | ···················· | ···················· | ···················· | ···················· | NNNNNNNNNNNNNNNNNNNN | NNNNNNNNNNNNNNNNNNNN | 1 |
| U | 4152 | hCoV-19/Belgium/ULG-10664/2020\|EPI_ISL_681265\|2020 | ···················· | ···················· | ···················· | ···················· | NNNNNNNNNNNNNNNNNNNN | NNNNNNNNNNNNNNNNNNNN | 1 |
| U | 4153 | hCoV-19/Belgium/ULG-10708/2020\|EPI_ISL_681266\|2020 | ···················· | ···················· | ···················· | ···················· | NNNNNNNNNNNNNNNNNNNN | NNNNNNNNNNNNNNNNNNNN | 1 |
| U | 4581 | hCoV-19/Saudi Arabia/KAUST-MADINAH1130/2020\|EPI_IS | ···················· | ···················· | ···················· | ···················· | ·······NNNNNNNNNNNNN | ···················· | 1 |
| U | 4595 | hCoV-19/Wales/PHWC-48FC2F/2020\|EPI_ISL_681204\|2020 | ···················· | ···················· | ···················· | ···················· | NNNNNNNNNNNNNNNNNNNN | ···················· | 1 |
| U | 4596 | hCoV-19/Wales/PHWC-48FC3E/2020\|EPI_ISL_681205\|2020 | ···················· | ···················· | ···················· | ···················· | NNNNNNNNNNNNNNNNNNNN | NNNNNNNNNNNNNNNNNNNN | 1 |
| U | 4597 | hCoV-19/Wales/PHWC-48FC4D/2020\|EPI_ISL_681206\|2020 | ···················· | ···················· | ···················· | ···················· | NNNNNNNNNNNNNNNNNNNN | NNNNNNNNNNNNNNNNNNNN | 1 |
| U | 4659 | hCoV-19/Wales/PHWC-48F551/2020\|EPI_ISL_681109\|2020 | ···················· | ···················· | ···················· | ···················· | NNNNNNNNNNNNNNNNNNNN | NNNNNNNNNNNNNNNNNNNN | 1 |
| U | 4663 | hCoV-19/Wales/PHWC-48F5E8/2020\|EPI_ISL_681115\|2020 | ···················· | ···················· | ···················· | ···················· | NNNNNNNNNNNNNNNNNNNN | NNNNNNNNNNNNNNNNNNNN | 1 |
| U | 4664 | hCoV-19/Wales/PHWC-48F5F7/2020\|EPI_ISL_681116\|2020 | ···················· | ···················· | ···················· | ···················· | NNNNNNNNNNNNNNNNNNNN | NNNNNNNNNNNNNNNNNNNN | 1 |
| U | 4682 | hCoV-19/Wales/PHWC-48F7C4/2020\|EPI_ISL_681139\|2020 | ···················· | ···················· | ···················· | ···················· | NNNNNNNNNNNNNNNNNNNN | NNNNNNNNNNNNNNNNNNNN | 1 |
| U | 4686 | hCoV-19/Wales/PHWC-48F83A/2020\|EPI_ISL_681146\|2020 | ···················· | ···················· | ···················· | ···················· | ··NNNNN············· | ···················· | 1 |
| U | 5085 | hCoV-19/Wales/PHWC-48F991/2020\|EPI_ISL_681167\|2020 | ···················· | ···················· | ···················· | ···················· | NNNNNNNNNNNNNNNNNNNN | ···················· | 1 |
| U | 5086 | hCoV-19/Wales/PHWC-48F9A0/2020\|EPI_ISL_681168\|2020 | ···················· | ···················· | ···················· | ···················· | NNNNNNNNNNNNNNNNNNNN | NNNNNNNNNNNNNNNNNNNN | 1 |
| U | 5088 | hCoV-19/Wales/PHWC-48F9DD/2020\|EPI_ISL_681170\|2020 | ···················· | ···················· | ···················· | ···················· | NNNNNNNNNNNNNNNNNNNN | NNNNNNNNNNNNNNNNNNNN | 1 |
| U | 5090 | hCoV-19/Wales/PHWC-48FA07/2020\|EPI_ISL_681173\|2020 | ···················· | ···················· | ···················· | ···················· | NNNNNNNNNNNNNNNNNNNN | ···················· | 1 |
| U | 5091 | hCoV-19/Wales/PHWC-48FA16/2020\|EPI_ISL_681174\|2020 | ···················· | ···················· | ···················· | ···················· | NNNNNNNNNNNNNNNNNNNN | NNNNNNNNNNNNNNNNNNNN | 1 |
| U | 5097 | hCoV-19/Wales/PHWC-48FAF8/2020\|EPI_ISL_681187\|2020 | ···················· | ···················· | ···················· | ···················· | NNNNNNNNNNNNNNNNNNNN | NNNNNNNNNNNNNNNNNNNN | 1 |
| U | 5098 | hCoV-19/Wales/PHWC-48FB04/2020\|EPI_ISL_681188\|2020 | ···················· | ···················· | ···················· | ···················· | ···················· | NNNNNN·············· | 1 |
| U | 5099 | hCoV-19/Wales/PHWC-48FB31/2020\|EPI_ISL_681190\|2020 | ···················· | ···················· | ···················· | ···················· | NNNNNNNNNNNNNNNNNNNN | NNNNNNNNNNNNNNNNNNNN | 1 |
| U | 5102 | hCoV-19/Wales/PHWC-48FB6E/2020\|EPI_ISL_681193\|2020 | ···················· | ···················· | ···················· | ···················· | ···················· | NNNNNNNNNNNNNNNNNNNN | 1 |
| U | 5106 | hCoV-19/Wales/PHWC-48FBAA/2020\|EPI_ISL_681197\|2020 | ···················· | ···················· | ···················· | ···················· | NNNNNNNNNNNNNNNNNNNN | NNNNNNNNNNNNNNNNNNNN | 1 |
| U | 5218 | hCoV-19/Wales/PHWC-48EDF0/2020\|EPI_ISL_681008\|2020 | ···················· | ···················· | ···················· | ···················· | NNNNNNNNNNNNNNNNNNNN | NNNNNNNNNNNNNNNNNNNN | 1 |
| U | 5222 | hCoV-19/Wales/PHWC-48EE57/2020\|EPI_ISL_681013\|2020 | ···················· | ···················· | ···················· | ···················· | NNNNNNNNNNNNNNNNNNNN | NNNNNNNNNNNNNNNNNNN· | 1 |
| U | 5224 | hCoV-19/Wales/PHWC-48EE75/2020\|EPI_ISL_681015\|2020 | ···················· | ···················· | ···················· | ···················· | NNNNNNNNNNNNNNNNNNNN | ···················· | 1 |
| U | 5227 | hCoV-19/Wales/PHWC-48EEFD/2020\|EPI_ISL_681020\|2020 | ···················· | ···················· | ···················· | ···················· | NNNNNNNNNNNNNNNNNNNN | NNNNNNNNNNNNNNNNNNNN | 1 |
| U | 5228 | hCoV-19/Wales/PHWC-48EF09/2020\|EPI_ISL_681021\|2020 | ···················· | ···················· | ···················· | ···················· | NNNNNNNNNNNNNNNNNNNN | NNNNNNNNNNNNNNNNNNNN | 1 |
| U | 5231 | hCoV-19/Wales/PHWC-48EFCD/2020\|EPI_ISL_681031\|2020 | ···················· | ···················· | ···················· | ···················· | NNNNNNNNNNNNNNNNNNNN | NNNNNNNNNNNNNNNNNNNN | 1 |
| U | 5233 | hCoV-19/Wales/PHWC-48EFEB/2020\|EPI_ISL_681033\|2020 | ···················· | ···················· | ···················· | ···················· | NNNNNNNNNNNNNNNNNNNN | NNN················· | 1 |
| U | 5238 | hCoV-19/Wales/PHWC-48F041/2020\|EPI_ISL_681038\|2020 | ···················· | ···················· | ···················· | ···················· | NNNNNNNNNNNNNNNNNNNN | ···················· | 1 |
| U | 5304 | hCoV-19/Spain/CT-IBV-97010161/2020\|EPI_ISL_683621\| | ···················· | ···················· | ···················· | ···················· | NNNNNNNNNNNNNNNNNNNN | NNNNNNNNNNNNNNNNNNNN | 1 |
| U | 5305 | hCoV-19/Spain/CT-IBV-97010117/2020\|EPI_ISL_683623\| | ···················· | ···················· | ···················· | ···················· | NNNNNNNNNNNNNNNNNNNN | NNNNNNNNNNNNNNNNNNNN | 1 |
| U | 5306 | hCoV-19/Spain/CT-IBV-97009572/2020\|EPI_ISL_683624\| | ···················· | ···················· | ···················· | ···················· | NNNNNNNNNNNNNNNNNNNN | NNNNNNNNNNNNNNNNNNNN | 1 |
| U | 5307 | hCoV-19/Spain/CT-IBV-97009588/2020\|EPI_ISL_683626\| | ···················· | ···················· | ···················· | ···················· | NNNNNNNNNNNNNNNNNNNN | NNNNNNNNNNNNNNNNNNNN | 1 |
| U | 5308 | hCoV-19/Spain/CT-IBV-97010155/2020\|EPI_ISL_683627\| | ···················· | ···················· | ···················· | ···················· | NNNNNNNNNNNNNNNNNNNN | NNNNNNNNNNNNNNNNNNNN | 1 |
| U | 5309 | hCoV-19/Spain/CT-IBV-97009584/2020\|EPI_ISL_683628\| | ···················· | ···················· | ···················· | ···················· | NNNNNNNNNNNNNNNNNNNN | NNNNNNNNNNNNNNNNNNNN | 1 |
| U | 5310 | hCoV-19/Spain/CT-IBV-97009580/2020\|EPI_ISL_683634\| | ···················· | ···················· | ···················· | ···················· | NNNNNNNNNNNNNNNNNNNN | NNNNNNNNNNNNNNNNNNNN | 1 |
| U | 5311 | hCoV-19/Spain/CT-IBV-97010114/2020\|EPI_ISL_683641\| | ···················· | ···················· | ···················· | ···················· | NNNNNNNNNNNNNNNNNNNN | NNNNNNNNNNNNNNNNNNNN | 1 |
| U | 5312 | hCoV-19/Spain/CT-IBV-97009578/2020\|EPI_ISL_683643\| | ···················· | ···················· | ···················· | ···················· | NNNNNNNNNNNNNNNNNNNN | NNNNNNNNNNNNNNNNNNNN | 1 |
| U | 5313 | hCoV-19/Spain/CT-IBV-97010135/2020\|EPI_ISL_683646\| | ···················· | ···················· | ···················· | ···················· | NNNNNNNNNNNNNNNNNNNN | NNNNNNNNNNNNNNNNNNNN | 1 |
| U | 5314 | hCoV-19/Spain/CT-IBV-97010112/2020\|EPI_ISL_683648\| | ···················· | ···················· | ···················· | ···················· | NNNNNNNNNNNNNNNNNNNN | NNNNNNNNNNNNNNNNNNNN | 1 |
| U | 5315 | hCoV-19/Spain/CT-IBV-97009574/2020\|EPI_ISL_683652\| | ···················· | ···················· | ···················· | ···················· | NNNNNNNNNNNNNNNNNNNN | NNNNNNNNNNNNNNNNNNNN | 1 |
| U | 5316 | hCoV-19/Spain/MD-IBV-97009994/2020\|EPI_ISL_683658\| | ···················· | ···················· | ···················· | ···················· | NNNNNNNNNNNNNNNNNNNN | NNNNNNNNNNNNNNNNNNNN | 1 |
| U | 5317 | hCoV-19/Spain/MD-IBV-97009995/2020\|EPI_ISL_683659\| | ···················· | ···················· | ···················· | ···················· | NNNNNNNNNNNNNNNNNNNN | NNNNNNNNNNNNNNNNNNNN | 1 |
| U | 5318 | hCoV-19/Spain/MD-IBV-97010000/2020\|EPI_ISL_683663\| | ···················· | ···················· | ···················· | ···················· | NNNNNNNNNNNNNNNNNNNN | NNNNNNNNNNNNNNNNNNNN | 1 |
| U | 5319 | hCoV-19/Spain/MD-IBV-97010003/2020\|EPI_ISL_683664\| | ·T·················· | ···················· | ···················· | ···················· | NNNNNNNNNNNNNNNNNNNN | NNNNNNNNNNNNNNNNNNNN | 1 |
| U | 5320 | hCoV-19/Spain/MD-IBV-97010008/2020\|EPI_ISL_683669\| | ···················· | ···················· | ···················· | ···················· | NNNNNNNNNNNNNNNNNNNN | NNNNNNNNNNNNNNNNNNNN | 1 |
| U | 5321 | hCoV-19/Spain/MD-IBV-97010009/2020\|EPI_ISL_683670\| | ···················· | ···················· | ···················· | ···················· | NNNNNNNNNNNNNNNNNNNN | NNNNNNNNNNNNNNNNNNNN | 1 |
| U | 5322 | hCoV-19/Spain/MD-IBV-97010017/2020\|EPI_ISL_683672\| | ···················· | ···················· | ···················· | ···················· | NNNNNNNNNNNNNNNNNNNN | NNNNNNNNNNNNNNNNNNNN | 1 |
| U | 5323 | hCoV-19/Spain/MD-IBV-97010019/2020\|EPI_ISL_683673\| | ···················· | ···················· | ···················· | ···················· | NNNNNNNNNNNNNNNNNNNN | NNNNNNNNNNNNNNNNNNNN | 1 |
| U | 5324 | hCoV-19/Spain/MD-IBV-97010020/2020\|EPI_ISL_683674\| | ···················· | ···················· | ···················· | ···················· | NNNNNNNNNNNNNNNNNNNN | NNNNNNNNNNNNNNNNNNNN | 1 |
| U | 5524 | hCoV-19/Hungary/US-55383w/2020\|EPI_ISL_677770\|2020 | ···················· | ···················· | ···················· | ···················· | NNNNNNNNNNNNNNNNNNNN | NNNNNNNNNNNNNNNNNNNN | 1 |
| U | 5525 | hCoV-19/Hungary/US-55426w/2020\|EPI_ISL_677771\|2020 | ···················· | ···················· | ···················· | ···················· | NNNNNNNNNNNNNNNNNNNN | NNNNNNNNNNNNNNNNNNNN | 1 |
| U | 5532 | hCoV-19/Hungary/US-57474w/2020\|EPI_ISL_677781\|2020 | ···················· | ···················· | ···················· | ···················· | NNNNNNNNNNNNNNNNNNNN | NNNNNNNNNNNNNNNNNNNN | 1 |
| U | 5535 | hCoV-19/Hungary/US-57510w/2020\|EPI_ISL_677784\|2020 | ···················· | ···················· | ···················· | ···················· | NNNNNNNNNNNNNNNNNNNN | NNNNNNNNNNNNNNNNNNNN | 1 |
| U | 5566 | hCoV-19/Hungary/US-63536w/2020\|EPI_ISL_677819\|2020 | ···················· | ···················· | ···················· | ···················· | NNNNNNNNNNNNNNNNNNNN | NNNNNNNNNNNNNNNNNNNN | 1 |
| U | 5575 | hCoV-19/Wales/PHWC-48F13F/2020\|EPI_ISL_681048\|2020 | ···················· | ···················· | ···················· | ···················· | NNNNNNNNNNNN········ | NNNNNNNNNNNNNNN····· | 1 |
| U | 5576 | hCoV-19/Wales/PHWC-48F15D/2020\|EPI_ISL_681050\|2020 | ···················· | ···················· | ···················· | ···················· | NNNNNNNNNNNNNNNN···· | ···················· | 1 |
| U | 5579 | hCoV-19/Wales/PHWC-48F199/2020\|EPI_ISL_681053\|2020 | ···················· | ···················· | ···················· | ···················· | NNNNNNNNNNNNNNNNNNNN | ···················· | 1 |
| U | 5596 | hCoV-19/Wales/PHWC-48F30C/2020\|EPI_ISL_681075\|2020 | ···················· | ···················· | ···················· | ···················· | ········NNNN········ | NNNNNNNNNNNNNNN····· | 1 |
| U | 5601 | hCoV-19/Wales/PHWC-48F375/2020\|EPI_ISL_681081\|2020 | ···················· | ···················· | ···················· | ···················· | NNNNNNNNNNNNNNNNNNNN | ···················· | 1 |
| U | 5614 | hCoV-19/Wales/PHWC-48F472/2020\|EPI_ISL_681096\|2020 | ···················· | ···················· | ···················· | ···················· | ···················· | NNNNN··············· | 1 |
| U | 5616 | hCoV-19/Wales/PHWC-48F490/2020\|EPI_ISL_681098\|2020 | ···················· | ···················· | ···················· | ···················· | NNNNNNNNNNNNNNNNNNNN | NNNNNNNNNNNNNNNNNNNN | 1 |
| U | 5847 | hCoV-19/Scotland/EDB10112/2020\|EPI_ISL_680474\|2020 | ···················· | ···················· | ···················· | ···················· | NNNNNNNNNNNNNNNNNNNN | NNNNNNNNNNNNNNNNNNNN | 1 |
| U | 5857 | hCoV-19/Scotland/EDB10150/2020\|EPI_ISL_680488\|2020 | ···················· | ···················· | ···················· | ···················· | NNNNNNNNNNNNNNNNNNNN | NNNNNNNNNNNNNNNNNNNN | 1 |
| U | 5868 | hCoV-19/Spain/CT-IBV-97009571/2020\|EPI_ISL_683601\| | ···················· | ···················· | ···················· | ···················· | NNNNNNNNNNNNNNNNNNNN | NNNNNNNNNNNNNNNNNNNN | 1 |
| U | 5869 | hCoV-19/Spain/CT-IBV-97009575/2020\|EPI_ISL_683603\| | ···················· | ···················· | ···················· | ···················· | NNNNNNNNNNNNNNNNNNNN | NNNNNNNNNNNNNNNNNNNN | 1 |
| U | 5870 | hCoV-19/Spain/CT-IBV-97009577/2020\|EPI_ISL_683604\| | ···················· | ···················· | ···················· | ···················· | NNNNNNNNNNNNNNNNNNNN | NNNNNNNNNNNNNNNNNNNN | 1 |
| U | 5871 | hCoV-19/Spain/CT-IBV-97010108/2020\|EPI_ISL_683606\| | ···················· | ···················· | ···················· | ···················· | NNNNNNNNNNNNNNNNNNNN | NNNNNNNNNNNNNNNNNNNN | 1 |
| U | 5872 | hCoV-19/Spain/CT-IBV-97010109/2020\|EPI_ISL_683607\| | ···················· | ···················· | ···················· | ···················· | NNNNNNNNNNNNNNNNNNNN | NNNNNNNNNNNNNNNNNNNN | 1 |
| U | 5873 | hCoV-19/Spain/CT-IBV-97010116/2020\|EPI_ISL_683608\| | ···················· | ···················· | ···················· | ···················· | NNNNNNNNNNNNNNNNNNNN | NNNNNNNNNNNNNNNNNNNN | 1 |
| U | 5874 | hCoV-19/Spain/CT-IBV-97010132/2020\|EPI_ISL_683610\| | ···················· | ···················· | ···················· | ···················· | NNNNNNNNNNNNNNNNNNNN | NNNNNNNNNNNNNNNNNNNN | 1 |
| U | 5875 | hCoV-19/Spain/CT-IBV-97010142/2020\|EPI_ISL_683612\| | ···················· | ···················· | ···················· | ···················· | NNNNNNNNNNNNNNNNNNNN | NNNNNNNNNNNNNNNNNNNN | 1 |
| U | 5876 | hCoV-19/Spain/CT-IBV-97010143/2020\|EPI_ISL_683613\| | ···················· | ···················· | ···················· | ···················· | NNNNNNNNNNNNNNNNNNNN | NNNNNNNNNNNNNNNNNNNN | 1 |
| U | 5877 | hCoV-19/Spain/CT-IBV-97010147/2020\|EPI_ISL_683616\| | ···················· | ···················· | ···················· | ···················· | NNNNNNNNNNNNNNNNNNNN | NNNNNNNNNNNNNNNNNNNN | 1 |
| U | 5878 | hCoV-19/Spain/CT-IBV-97010153/2020\|EPI_ISL_683619\| | ···················· | ···················· | ···················· | ···················· | NNNNNNNNNNNNNNNNNNNN | NNNNNNNNNNNNNNNNNNNN | 1 |
| U | 6082 | hCoV-19/Northern Ireland/NIRE-234784/2020\|EPI_ISL_ | ···················· | ···················· | ···················· | ···················· | NNNN················ | NNNNNNNNNNNNNNN····· | 1 |
| U | 6097 | hCoV-19/Scotland/EDB10050/2020\|EPI_ISL_680463\|2020 | ···················· | ···················· | ···················· | ···················· | NNNNNNNNNNNNNNNNNNNN | NNNNNNNNNNNNNNNNNNNN | 1 |

U 6098 hCoV-19/Scotland/EDB10077/2020|EPI_ISL_680464|2020 ···················· ···················· ···················· ···················· NNNNNNNNNNNNNNNNNNNN NNNNNNNNNNNNNNNNNNNN 1
U 6099 hCoV-19/Scotland/EDB10081/2020|EPI_ISL_680465|2020 ···················· ···················· ···················· ···················· NNNNNNNNNNNNNNNNNNNN NNNNNNNNNNNNNNNNNNNN 1
U 6102 hCoV-19/Scotland/EDB10092/2020|EPI_ISL_680468|2020 ···················· ···················· ···················· ···················· NNNNNNNNNNNNNNNNNNNN NNNNNNNNNNNNNNNNNNNN 1
U 6106 hCoV-19/Scotland/EDB10110/2020|EPI_ISL_680472|2020 ···················· ···················· ···················· ···················· NNNNNNNNNNNNNNNNNNNN NNNNNNNNNNNNNNNNNNNN 1
U 6261 hCoV-19/Saudi Arabia/KAUST-MAKKAH199/2020|EPI_ISL_ ···················· ···················· ···················· ···················· ········NNNNNNNNNNNN ···················· 1
U 6305 hCoV-19/England/LOND-1251539/2020|EPI_ISL_679409|2 ···················· ···················· ···················· ···················· NNNNNNNNNNNNNNNNNNNN NNNNNNNNNNNNNNNNNNNN 1
U 6307 hCoV-19/England/LOND-1251593/2020|EPI_ISL_679412|2 ···················· ···················· ···················· ···················· NNNNNNNNNNNNNNNNNNNN NNNNNNNNNNNNNNNNNNNN 1
U 6318 hCoV-19/England/LOND-12517AC/2020|EPI_ISL_679436|2 ···················· ···················· ···················· ···················· NNNNNNNNNNNNNNNNNNNN NNNNNNNNNNNNNNNNNNNN 1
U 6332 hCoV-19/Northern Ireland/NIRE-2342ED/2020|EPI_ISL_ ···················· ···················· ···················· ···················· NNNNNNNNNNNNN······· ···················· 1
U 6333 hCoV-19/Northern Ireland/NIRE-2342FC/2020|EPI_ISL_ ···················· ···················· ···················· ···················· ··········NNNNNN···· ···················· 1
U 6334 hCoV-19/Northern Ireland/NIRE-234335/2020|EPI_ISL_ ···················· ···················· ···················· ···················· NNNNNNNNNNNNNNNNNNNN ···················· 1
U 6336 hCoV-19/Northern Ireland/NIRE-234353/2020|EPI_ISL_ ···················· ···················· ···················· ···················· NNNNNNNNNNNNNNNNNNNN ···················· 1
U 6339 hCoV-19/Northern Ireland/NIRE-23439F/2020|EPI_ISL_ ···················· ···················· ···················· ···················· N·······NNNNNNN····· ···················· 1
U 6343 hCoV-19/Northern Ireland/NIRE-2343EA/2020|EPI_ISL_ ···················· ···················· ···················· ···················· NNNNNNNNNNNN········ ···················· 1
U 6346 hCoV-19/Northern Ireland/NIRE-234414/2020|EPI_ISL_ ···················· ···················· ···················· ···················· ···················· NNNNNNNNNNNNNNNNNNNN 1
U 6357 hCoV-19/Northern Ireland/NIRE-2344D8/2020|EPI_ISL_ ···················· ···················· ···················· ···················· ·NN··········NNNNNNN ···················· 1
U 6359 hCoV-19/Northern Ireland/NIRE-2344F6/2020|EPI_ISL_ ···················· ···················· ···················· ···················· NNNNNNNNNNNNNNNNNNNN ···················· 1
U 6368 hCoV-19/Northern Ireland/NIRE-23462D/2020|EPI_ISL_ ···················· ···················· ···················· ···················· NNNNNNNNNNNNNNNNNNNN NNNNNNNNNNNNNNN····· 1
U 6369 hCoV-19/Northern Ireland/NIRE-234696/2020|EPI_ISL_ ···················· ···················· ···················· ···················· NNNNNNNNNNNNNNNNNNNN ···················· 1
U 6370 hCoV-19/Northern Ireland/NIRE-2346A5/2020|EPI_ISL_ ···················· ···················· ···················· ···················· NNNNNNNNNNNNNNNNNNNN NNNNNNNNNNNNNNNNNNNN 1
U 6371 hCoV-19/Northern Ireland/NIRE-2346C3/2020|EPI_ISL_ ···················· ···················· ···················· ···················· NNNNNNNNNNNNNNNNNNNN NNNNNNNNNNNNNNN····· 1
U 6374 hCoV-19/Northern Ireland/NIRE-23470C/2020|EPI_ISL_ ···················· ···················· ···················· ···················· NNNNNNNNNNNNNNNNNNNN ···················· 1
U 6375 hCoV-19/Northern Ireland/NIRE-23471B/2020|EPI_ISL_ ···················· ···················· ···················· ···················· NNNNNNNNNNNNNNNNNNNN NNNNNNNNNNNNNNNNNNNN 1
U 6376 hCoV-19/Northern Ireland/NIRE-23472A/2020|EPI_ISL_ ···················· ···················· ···················· ···················· NNNNNNNNNNNNNNNNNNNN ···················· 1
U 6583 hCoV-19/Saudi Arabia/KAUST-MADINAH210/2020|EPI_ISL ···················· ···················· ···················· ···················· ·······NNNNNNNNNNNNN ···················· 1
U 6605 hCoV-19/Northern Ireland/NIRE-234010/2020|EPI_ISL_ ···················· ···················· ···················· ···················· NNNNNNNNNNNNNNNNNNNN NNNNNNNNN··········· 1
U 6606 hCoV-19/Northern Ireland/NIRE-23402F/2020|EPI_ISL_ ···················· ···················· ···················· ···················· NNNN················ NNNNNNNNNNNNNNNNNNNN 1
U 6610 hCoV-19/Northern Ireland/NIRE-23406B/2020|EPI_ISL_ ···················· ···················· ···················· ···················· NNNNNNNNNNNNNNNNNNNN ···················· 1
U 6614 hCoV-19/Northern Ireland/NIRE-2340F2/2020|EPI_ISL_ ···················· ···················· ···················· ···················· ·············NNNNNNN NNNNNNNNNNNNNNNNNNNN 1
U 6628 hCoV-19/Chile/MA-CADIUMAG-8/2020|EPI_ISL_681674|20 ···················· ···················· ···················· ···················· NNNNNNNNNNNNNNNNNNNN NNNNNNNNNNNNNNNNNNNN 1
U 6629 hCoV-19/Chile/MA-CADIUMAG-9/2020|EPI_ISL_681675|20 ···················· ···················· ···················· ···················· NNNNNNNNNNNNNNNNNNNN NNNNNNNNNNNNNNNNNNNN 1
U 6743 hCoV-19/Northern Ireland/NIRE-232EEB/2020|EPI_ISL_ ···················· ···················· ···················· ···················· NNNNN··············· ···················· 1
U 6744 hCoV-19/Northern Ireland/NIRE-232F9D/2020|EPI_ISL_ ···················· ···················· ···················· ···················· NNNNNNNNNNNNNNNNNNNN ···················· 1
U 6746 hCoV-19/Northern Ireland/NIRE-232FCA/2020|EPI_ISL_ ···················· ···················· ···················· ···················· NNNNNNNNNNNNNNNNNNNN NNNNNNNNNNNNNNNNNNNN 1
U 6747 hCoV-19/Northern Ireland/NIRE-232FF7/2020|EPI_ISL_ ···················· ···················· ···················· ···················· NNNNNNNNNNNNNNNNNNNN NNNNNNNNNNNNNNNNNNNN 1
U 6748 hCoV-19/Northern Ireland/NIRE-233002/2020|EPI_ISL_ ···················· ···················· ···················· ···················· NNNN················ NNNNN··············· 1
U 6749 hCoV-19/Northern Ireland/NIRE-233011/2020|EPI_ISL_ ···················· ···················· ···················· ···················· ···················· NNNNNNNNNNNNNNN····· 1
U 6750 hCoV-19/Northern Ireland/NIRE-233020/2020|EPI_ISL_ ···················· ···················· ···················· ···················· NNNNNNNNNNNNNNNNNNNN ···················· 1
U 6751 hCoV-19/Northern Ireland/NIRE-23303F/2020|EPI_ISL_ ···················· ···················· ···················· ···················· ··NNNNNNNNNNNNNNNNNN ···················· 1
U 6752 hCoV-19/Northern Ireland/NIRE-23306C/2020|EPI_ISL_ ···················· ···················· ···················· ···················· NNNNNNNNNNNNN······· ···················· 1
U 6753 hCoV-19/Northern Ireland/NIRE-23307B/2020|EPI_ISL_ ···················· ···················· ···················· ···················· NNNNNNNNNNNNNNNNNNNN ·········NNNNNNNNNNN 1
U 6756 hCoV-19/Northern Ireland/NIRE-23314B/2020|EPI_ISL_ ···················· ···················· ···················· ···················· NNNNNNNNNNNNNNNNNNNN ···················· 1
U 6757 hCoV-19/Northern Ireland/NIRE-233178/2020|EPI_ISL_ ···················· ···················· ···················· ···················· NNNNNNNNNNNNNNNNNNNN NNNNNNNNNNNNNNNNNNNN 1
U 6758 hCoV-19/Northern Ireland/NIRE-233187/2020|EPI_ISL_ ···················· ···················· ···················· ···················· NNNNNNNNNNNNNNNNNNNN NNNNNNNNNNNNNNNNNNNN 1
U 6760 hCoV-19/Northern Ireland/NIRE-2331F0/2020|EPI_ISL_ ···················· ···················· ···················· ···················· NNNNNNNNNNNNNNNNNNNN NNNNNNNNNNNNNNNNNNNN 1
U 6762 hCoV-19/Northern Ireland/NIRE-23322A/2020|EPI_ISL_ ···················· ···················· ···················· ···················· NNNNNNNNNNNNNNNNNNNN NNNNNNNNNNNNNNNNNNNN 1
U 6763 hCoV-19/Northern Ireland/NIRE-233239/2020|EPI_ISL_ ···················· ···················· ···················· ···················· NNNNNNNNNNNNNNNNNNNN ···················· 1
U 6764 hCoV-19/Northern Ireland/NIRE-233248/2020|EPI_ISL_ ···················· ···················· ···················· ···················· NNNNN········NNNNNNN NNNNNNNNNNNNNNNNNNNN 1
U 6765 hCoV-19/Northern Ireland/NIRE-233257/2020|EPI_ISL_ ···················· ···················· ···················· ···················· NNNNNNNNNNNNNNNNNNNN ···················· 1
U 6766 hCoV-19/Northern Ireland/NIRE-233266/2020|EPI_ISL_ ···················· ···················· ···················· ···················· NNNNNNNNNNNNNNNNNNNN ···················· 1
U 6768 hCoV-19/Northern Ireland/NIRE-2332A2/2020|EPI_ISL_ ···················· ···················· ···················· ···················· NNNNNNNNNNNNNNNNNNNN NNNNNNNNNNNNNNNN···· 1
U 6769 hCoV-19/Northern Ireland/NIRE-2332DF/2020|EPI_ISL_ ···················· ···················· ···················· ···················· NNNNNNNNNNNNNNNNNNNN ···················· 1
U 6770 hCoV-19/Northern Ireland/NIRE-2332EE/2020|EPI_ISL_ ···················· ···················· ···················· ···················· NNNNNNNNNNNNNNN····· NNNNNNNNNNNNNNNNNNNN 1
U 6771 hCoV-19/Northern Ireland/NIRE-2332FD/2020|EPI_ISL_ ···················· ···················· ···················· ···················· NNNNNNNNNNNNNNNNNNNN NNNNNNNNNNNNNNNNNNNN 1
U 6772 hCoV-19/Northern Ireland/NIRE-233327/2020|EPI_ISL_ ···················· ···················· ···················· ···················· NNNNNNNNNNNNNNNNNNNN NNNNNNNNNNNNNNNNNNNN 1
U 6774 hCoV-19/Northern Ireland/NIRE-233345/2020|EPI_ISL_ ···················· ···················· ···················· ···················· NNNNNNNNNNNNNNNNNNNN NNNNNNNNNNNNNNNNNNNN 1
U 6775 hCoV-19/Northern Ireland/NIRE-2333AF/2020|EPI_ISL_ ···················· ···················· ···················· ···················· NNNNNNNNNNNNNNNNNNNN NNNNNNNNNNNNNNNNNNNN 1
U 6776 hCoV-19/Northern Ireland/NIRE-2333BE/2020|EPI_ISL_ ···················· ···················· ···················· ···················· NNNNNNNNNNNNNNNNNNNN NNNN················ 1
U 6792 hCoV-19/Northern Ireland/NIRE-233FBA/2020|EPI_ISL_ ···················· ···················· ···················· ···················· NNNN················ ···················· 1
U 7150 hCoV-19/Northern Ireland/NIRE-10974D/2020|EPI_ISL_ ···················· ···················· ···················· ···················· NNNN················ ···················· 1
U 7613 hCoV-19/USA/NY-Wadsworth-103677-01/2020|EPI_ISL_67 ···················· ···················· ···················· ···················· NNNNNNN············· ···················· 1
U 7627 hCoV-19/USA/NY-Wadsworth-116619-01/2020|EPI_ISL_67 ···················· ···················· ···················· ···················· NNNNNNN············· ···················· 1
V 107 hCoV-19/Japan/PG-2101/2020|EPI_ISL_685706|2020-04 ···················· ···················· ···················· ···················· ···················· NNNNNNNNNNNNNNNNNNNN 1
V 242 hCoV-19/Japan/PG-5195/2020|EPI_ISL_690298|2020-08 ···················· ···················· ···················· ···················· ···················· NNNNNNNNNNNNNNNN···· 1
V 376 hCoV-19/Japan/PG-5145/2020|EPI_ISL_690252|2020-08 ···················· ···················· ···················· ···················· ···················· NNNNNNNNNNNNNNNNNNNN 1
V 385 hCoV-19/Japan/PG-5154/2020|EPI_ISL_690261|2020-08 ···················· ···················· ···················· ···················· ···················· NNNNNNNNNNNNNNNNNNNN 1
V 410 hCoV-19/Japan/PG-3424/2020|EPI_ISL_691536|2020-04 ···················· ···················· ···················· ···················· ···················· ················NNNN 1
V 411 hCoV-19/Japan/PG-3425/2020|EPI_ISL_691537|2020-04 ···················· ···················· ···················· ···················· NNNNNNNNNNNNNNNNNNNN NNNNNNNNNNNNNNNNNNNN 1
V 413 hCoV-19/Japan/PG-3428/2020|EPI_ISL_691539|2020-04 ···················· ···················· ···················· ···················· ···················· NNNNNNNNNNNNNNNNNNNN 1
V 635 hCoV-19/Japan/PG-7846/2020|EPI_ISL_686947|2020-08 ···················· ···················· ···················· ···················· ···················· NNNNNNNNNNNNNNNNNNNN 1
V 743 hCoV-19/Japan/PG-3382/2020|EPI_ISL_691497|2020-04 ···················· ···················· ···················· ···················· ···················· NNNNNNNNNNNNNNNNNNNN 1
V 745 hCoV-19/Japan/PG-3384/2020|EPI_ISL_691499|2020-04 ···················· ···················· ···················· ···················· ···················· NNNNNNNNNNNNNNNNNNNN 1
V 786 hCoV-19/Japan/PG-0719/2020|EPI_ISL_684329|2020-04 ···················· ···················· ···················· ···················· ···················· ··············NNNNNN 1
V 836 hCoV-19/Japan/PG-2039/2020|EPI_ISL_685644|2020-05 ···················· ···················· ···················· ···················· ···················· ····NNNNNNNNNNNNNNNN 1
V 852 hCoV-19/Japan/PG-2055/2020|EPI_ISL_685660|2020-04 ···················· ···················· ···················· ···················· ···················· NNNNNNNNNNNNNNNNNNNN 1
V 859 hCoV-19/Japan/PG-2062/2020|EPI_ISL_685667|2020-04 ···················· ···················· ···················· ···················· ···················· NNNNNNNNNNNNNNNNNNNN 1
V 860 hCoV-19/Japan/PG-2063/2020|EPI_ISL_685668|2020-04 ···················· ···················· ···················· ···················· ···················· NNNNNNNNNNNNNNNNNNNN 1
V 863 hCoV-19/Japan/PG-2066/2020|EPI_ISL_685671|2020-04 ···················· ···················· ···················· ···················· NNNNNNNNNNNNNNNNNNNN NNNNNNNNNNNNNNNNNNNN 1
V 873 hCoV-19/Japan/PG-2076/2020|EPI_ISL_685681|2020-04 ···················· ···················· ···················· ···················· ···················· NNNNNNNNNNNNNNNNNNNN 1
V 875 hCoV-19/Japan/PG-2078/2020|EPI_ISL_685683|2020-04 ···················· ···················· ···················· ···················· ···················· NNNNNNNNNNNNNNNNNNNN 1

| | | | | | | | | | |
|---|---|---|---|---|---|---|---|---|---|
| V | 876 | hCoV-19/Japan/PG-2079/2020\|EPI_ISL_685684\|2020-04 | ···················· | ···················· | ···················· | ···················· | ···················· | NNNNNNN············· | 1 |
| V | 978 | hCoV-19/Japan/PG-3313/2020\|EPI_ISL_691428\|2020-04 | ···················· | ···················· | ···················· | ···················· | ···················· | NNNNNNNNNNNNNNNNNNNN | 1 |
| V | 981 | hCoV-19/Japan/PG-3316/2020\|EPI_ISL_691431\|2020-04 | ···················· | ···················· | ···················· | ···················· | ···················· | ·····NNNNN·········· | 1 |
| V | 1049 | hCoV-19/Japan/PG-1908/2020\|EPI_ISL_685514\|2020-04 | ···················· | ···················· | ···················· | ···················· | ···················· | NNN··NNNNNNNNNN····· | 1 |
| V | 1174 | hCoV-19/Japan/PG-5031/2020\|EPI_ISL_690142\|2020-07 | ···················· | NNNNNNNNNNNNNNNNNNNN | ···················· | ···················· | ···················· | ···················· | 1 |
| V | 1187 | hCoV-19/Japan/PG-3325/2020\|EPI_ISL_691440\|2020-04 | ···················· | ···················· | ···················· | ···················· | ···················· | N·NNNNNNNN·········· | 1 |
| V | 1249 | hCoV-19/Japan/PG-4495/2020\|EPI_ISL_692718\|2020-07 | ···················· | ···················· | ···················· | ···················· | ···················· | NNNNNNNNNNNNNNNNNNNN | 1 |
| V | 1344 | hCoV-19/Japan/PG-1979/2020\|EPI_ISL_685585\|2020-02 | ···················· | ···················· | ···················· | ···················· | NNNNNNNNNNNNNNNNNNNN | NNNNNNNNNNNNNNNNNNNN | 1 |
| V | 1457 | hCoV-19/Japan/PG-4919/2020\|EPI_ISL_690039\|2020-08 | ···················· | ···················· | ···················· | ···················· | ···················· | NNNNNNNNNNNNNNNNNNNN | 1 |
| V | 1536 | hCoV-19/Japan/PG-4476/2020\|EPI_ISL_692699\|2020-07 | ···················· | ···················· | ···················· | ···················· | ···················· | NNNNNNNNNNNNNNNNNNNN | 1 |
| V | 1720 | hCoV-19/Japan/PG-4388/2020\|EPI_ISL_692615\|2020-07 | ···················· | ···················· | ···················· | ···················· | ···················· | ······NNNNNNNNNNNNNN | 1 |
| V | 1730 | hCoV-19/Japan/PG-4399/2020\|EPI_ISL_692625\|2020-07 | ···················· | ···················· | ···················· | ···················· | ···················· | NNNNNNNNNNNNNNNNNNNN | 1 |
| V | 1736 | hCoV-19/Japan/PG-4405/2020\|EPI_ISL_692631\|2020-07 | ···················· | ···················· | ···················· | ···················· | NNNNNNNNNNNNNNNNNNNN | NNNNNNNNNNNNNNNNNNNN | 1 |
| V | 2001 | hCoV-19/Japan/PG-4459/2020\|EPI_ISL_692682\|2020-07 | ···················· | ···················· | ···················· | ···················· | ···················· | NNNNNNNNNNNNNNNNNNNN | 1 |
| V | 2700 | hCoV-19/Japan/PG-4548/2020\|EPI_ISL_689686\|2020 | ···················· | ···················· | ···················· | ···················· | NNNNNNNNNNNNNNNNNNNN | NNNNNNNNNNNNNNNNNNNN | 1 |
| V | 2702 | hCoV-19/Japan/PG-4550/2020\|EPI_ISL_689688\|2020 | ···················· | ···················· | ···················· | ···················· | ···················· | NNNNNNN············· | 1 |
| V | 3231 | hCoV-19/Japan/PG-4552/2020\|EPI_ISL_689690\|2020 | ···················· | ···················· | ···················· | ···················· | ···················· | ··············NNNNNN | 1 |
| V | 3438 | hCoV-19/Japan/PG-6436/2020\|EPI_ISL_689542\|2020-07 | ···················· | ···················· | ···················· | ···················· | ···················· | ·········NNNNNNN···· | 1 |
| V | 3487 | hCoV-19/Japan/PG-5461/2020\|EPI_ISL_690564\|2020-08 | ···················· | ···················· | ···················· | ···················· | ···················· | NNNNNNNNNNNNNNNNNNNN | 1 |
| V | 3631 | hCoV-19/Japan/PG-2354/2020\|EPI_ISL_685959\|2020-04 | ···················· | ···················· | ···················· | ···················· | NNNNNNNNN··········· | NNNNNNNNNNNNNNNNNNNN | 1 |
| V | 3636 | hCoV-19/Japan/PG-2359/2020\|EPI_ISL_685964\|2020-05 | ···················· | ···················· | ···················· | ···················· | ···················· | NNNNNNNNNNNNNNNNNNNN | 1 |
| V | 3654 | hCoV-19/Japan/PG-2377/2020\|EPI_ISL_685982\|2020-05 | ···················· | ···················· | ···················· | ···················· | ········NNNNNNNNNNNN | ···················· | 1 |
| V | 3746 | hCoV-19/Japan/PG-5410/2020\|EPI_ISL_690513\|2020-08 | ···················· | ···················· | ···················· | ···················· | NNNNNNNNNNNNNNNNNNNN | NNNNNNNNNNNNNNNNNNNN | 1 |
| V | 3748 | hCoV-19/Japan/PG-5413/2020\|EPI_ISL_690516\|2020-08 | ···················· | ···················· | ···················· | ···················· | NNNNNNNNNNNNNNNNNNNN | NNNNNNNNNNNNNNNNNNNN | 1 |
| V | 3925 | hCoV-19/Japan/PG-2349/2020\|EPI_ISL_685954\|2020-04 | ···················· | ···················· | ···················· | ···················· | ···················· | NNNNNNNN············ | 1 |
| V | 3929 | hCoV-19/Japan/PG-2353/2020\|EPI_ISL_685958\|2020-04 | ···················· | ···················· | ···················· | ···················· | ···················· | NNNNNNNNNNN········· | 1 |
| V | 3964 | hCoV-19/Japan/PG-6305/2020\|EPI_ISL_689416\|2020-08 | ···················· | ···················· | ···················· | ···················· | ···················· | ··············NNNNNN | 1 |
| V | 3966 | hCoV-19/Japan/PG-6307/2020\|EPI_ISL_689418\|2020-08 | ···················· | ···················· | ···················· | ···················· | NNNNNNNNN··········· | NNNNNNNNNNNNNNN····· | 1 |
| V | 3979 | hCoV-19/Japan/PG-6320/2020\|EPI_ISL_689431\|2020-08 | ···················· | ···················· | ···················· | ···················· | ···················· | NNNNNNNNNNNNNNNNNNNN | 1 |
| V | 4123 | hCoV-19/Japan/PG-2296/2020\|EPI_ISL_685901\|2020-04 | ···················· | ···················· | ···················· | ···················· | NNNNNNNNNNNNNNNNNNNN | NNNNNNNNNNNNNNNNNNNN | 1 |
| V | 4139 | hCoV-19/Japan/PG-2312/2020\|EPI_ISL_685917\|2020-05 | ···················· | ···················· | ···················· | ···················· | ···················· | NNNNN···NNNNNNNNNNNN | 1 |
| V | 4154 | hCoV-19/Japan/PG-7025/2020\|EPI_ISL_688130\|2020-08 | ···················· | ···················· | ···················· | ···················· | ···················· | ··NNNNN············· | 1 |
| V | 4173 | hCoV-19/Japan/PG-7044/2020\|EPI_ISL_688149\|2020-08 | ···················· | ···················· | ···················· | ···················· | ···················· | ······NNNNNNNNNNNNNN | 1 |
| V | 4210 | hCoV-19/Japan/PG-7081/2020\|EPI_ISL_688186\|2020-08 | ···················· | ···················· | ···················· | ···················· | ···················· | NNNNNNNNNNNNNNNNNNNN | 1 |
| V | 4295 | hCoV-19/Japan/PG-5317/2020\|EPI_ISL_690420\|2020-08 | ···················· | ···················· | ···················· | ···················· | ···················· | NNNNNNNNNNNN··NNNNNN | 1 |
| V | 4297 | hCoV-19/Japan/PG-5319/2020\|EPI_ISL_690422\|2020-08 | ···················· | ···················· | ···················· | ···················· | ···NNNNNNNNNNNNNNNNN | ········NNNNNNNNNNNN | 1 |
| V | 4776 | hCoV-19/Japan/PG-5284/2020\|EPI_ISL_690387\|2020-07 | ···················· | ···················· | ···················· | ···················· | ···················· | NNNNNNNNNNNNNNNNNNNN | 1 |
| V | 4794 | hCoV-19/Spain/IB-IBV-99010754/2020\|EPI_ISL_691661\| | ···················· | ···················· | ···················· | ···················· | ·NNNNNN············· | ···················· | 1 |
| V | 4811 | hCoV-19/Spain/IB-IBV-99010804/2020\|EPI_ISL_691678\| | ···················· | ···················· | ···················· | ···················· | NNNNNNNNNNNNNNNNNNNN | NNNNNNNNNNNNNNNNNNNN | 1 |
| V | 4812 | hCoV-19/Spain/IB-IBV-99010808/2020\|EPI_ISL_691679\| | ···················· | ···················· | ···················· | ···················· | ··NNNNN············· | ···················· | 1 |
| V | 4968 | hCoV-19/Japan/PG-5752/2020\|EPI_ISL_688871\|2020-08 | ···················· | NNNNNNNNNNNNNNNNNNNN | ···················· | ···················· | ···················· | ···················· | 1 |
| V | 5002 | hCoV-19/Japan/PG-5202/2020\|EPI_ISL_690305\|2020-08 | ···················· | ···················· | ···················· | ···················· | NNNNNNNNNNNNNNNNNNNN | NNNNNNNNNNNNNNNNNNNN | 1 |
| V | 5111 | hCoV-19/Thailand/Bangkok-CONI-0254/2020\|EPI_ISL_69 | ···················· | ···················· | ···················· | ···················· | ·NNNNNNNNNNNNNNNNNNN | NNNNNNNNNNNNNNNNNNNN | 1 |
| V | 5112 | hCoV-19/Thailand/Bangkok-CONI-0256/2020\|EPI_ISL_69 | ···················· | ···················· | ···················· | ···················· | NNNNNNNNNNNNNNNNNNNN | NNNNNNNNNNNNNNNNNNNN | 1 |
| V | 5113 | hCoV-19/Thailand/Bangkok-CONI-0257/2020\|EPI_ISL_69 | ···················· | ···················· | ···················· | ···················· | ·NNNNNNNNNNNNNNNNNNN | NNNNNNNNNNNNNNNNNNNN | 1 |
| V | 5114 | hCoV-19/Thailand/Bangkok-CONI-0258/2020\|EPI_ISL_69 | ···················· | ···················· | ···················· | ···················· | ··NNNNNNNNNNNNNNNNNN | ·················NNN | 1 |
| V | 5115 | hCoV-19/Thailand/Bangkok-CONI-0260/2020\|EPI_ISL_69 | ···················· | ···················· | ···················· | ···················· | ··NNNNNNNNNNNNNNNNNN | ···················· | 1 |
| V | 5188 | hCoV-19/Japan/PG-8365/2020\|EPI_ISL_687464\|2020-07 | ···················· | ···················· | ···················· | ···················· | ···················· | NNNNNNNNNNNNNNNNNNNN | 1 |
| V | 5197 | hCoV-19/Japan/PG-8374/2020\|EPI_ISL_687473\|2020-07 | ···················· | ···················· | ···················· | ···················· | ···················· | NN·······NNNNNNNNNNN | 1 |
| V | 5335 | hCoV-19/Thailand/Bangkok-CONI-0261/2020\|EPI_ISL_69 | ···················· | ···················· | ···················· | ···················· | ·NNNNNNNNNNNNNNNNNNN | NNNNNNNNNNNNNNNN···· | 1 |
| V | 5336 | hCoV-19/Thailand/Bangkok-CONI-0262/2020\|EPI_ISL_69 | ···················· | ···················· | ···················· | ···················· | ·NNNNNNNNNNNNNNNNNNN | NNNNNNNNNNNNNNNNNNNN | 1 |
| V | 5337 | hCoV-19/Thailand/Bangkok-CONI-0263/2020\|EPI_ISL_69 | ···················· | ···················· | ···················· | ···················· | ··NNNNNNNNNNNNNNNNNN | NNNNNNNNNNNNNNNNNNNN | 1 |
| V | 5338 | hCoV-19/Thailand/Bangkok-CONI-0264/2020\|EPI_ISL_69 | ···················· | ···················· | ···················· | ···················· | ·NNNNNNNNNNNNNNNNNNN | NNNNNNNNNNNNNNNNNNNN | 1 |
| V | 5339 | hCoV-19/Thailand/Bangkok-CONI-0265/2020\|EPI_ISL_69 | ···················· | ···················· | ···················· | ···················· | ·NNNNNNNNNNNNNNNNNNN | NNNNNNNNNNNNNNNNNNNN | 1 |
| V | 5340 | hCoV-19/Thailand/Bangkok-CONI-0267/2020\|EPI_ISL_69 | ···················· | ···················· | ···················· | ···················· | ···················· | NNNNNNNNNNNNNNNNNNNN | 1 |
| V | 5341 | hCoV-19/Thailand/Bangkok-CONI-0269/2020\|EPI_ISL_69 | ···················· | ···················· | ···················· | ···················· | ·NNNNNNNNNNNNNNNNNNN | NNNNNNNNNNNNNNNNNNNN | 1 |
| V | 5342 | hCoV-19/Thailand/Bangkok-CONI-0270/2020\|EPI_ISL_69 | ···················· | ···················· | ···················· | ···················· | ·NNNNNNNNNNNNNNNNNNN | NNNNNNNNNNNNNNNNNNNN | 1 |
| V | 5343 | hCoV-19/Thailand/Bangkok-CONI-0271/2020\|EPI_ISL_69 | ···················· | ···················· | ···················· | ···················· | NNNNNNNNNNNNNNNNNNNN | NNNNNNNNNNNNNNNNNNNN | 1 |
| V | 5344 | hCoV-19/Thailand/Bangkok-CONI-0272/2020\|EPI_ISL_69 | ···················· | ···················· | ···················· | ···················· | ·NNNNNNNNNNNNNNNNNNN | NNNNNNNNNNNNNNNNNNNN | 1 |
| V | 5345 | hCoV-19/Thailand/Bangkok-CONI-0273/2020\|EPI_ISL_69 | ···················· | ···················· | ···················· | ···················· | ·NNNNNNNNNNNNNNNNNNN | ···················· | 1 |
| V | 5347 | hCoV-19/Thailand/Bangkok-CONI-0275/2020\|EPI_ISL_69 | ···················· | ···················· | ···················· | ···················· | ··NNNNNNNNNNNNNNNNNN | NNNNNNNNNNNNNNNNNNNN | 1 |
| V | 5348 | hCoV-19/Thailand/Bangkok-CONI-0276/2020\|EPI_ISL_69 | ···················· | ···················· | ···················· | ···················· | ·NNNNNNNNNNNNNNNNNNN | NNNNNNNNNNNNNNNNNNNN | 1 |
| V | 5349 | hCoV-19/Thailand/Bangkok-CONI-0277/2020\|EPI_ISL_69 | ···················· | ···················· | ···················· | ···················· | ·NNNNNNNNNNNNNNNNNNN | NNNNNNNNNNNNNNNNNNNN | 1 |
| V | 5350 | hCoV-19/Thailand/Bangkok-CONI-0278/2020\|EPI_ISL_69 | ···················· | ···················· | ···················· | ···················· | ·NNNNNNNNNNNNNNNNNNN | NNNNNNNNNNNNNNNNNNNN | 1 |
| V | 5351 | hCoV-19/Thailand/Bangkok-CONI-0279/2020\|EPI_ISL_69 | ···················· | ···················· | ···················· | ···················· | ·NNNNNNNNNNNNNNNNNNN | NNNNNNNNNNNNNNNNNNNN | 1 |
| V | 5353 | hCoV-19/Thailand/Bangkok-CONI-0281/2020\|EPI_ISL_69 | ···················· | ···················· | ···················· | ···················· | NNNNNNNNNNNNNNNNNNNN | NNNNNNNNNNNNNNNNNNNN | 1 |
| V | 5354 | hCoV-19/Thailand/Bangkok-CONI-0283/2020\|EPI_ISL_69 | ···················· | ···················· | ···················· | ···················· | ·NNNNNNNNNNNNNNNNNNN | NNNNNNNNNNNNNNNNNNNN | 1 |
| V | 5355 | hCoV-19/Thailand/Bangkok-CONI-0289/2020\|EPI_ISL_69 | ···················· | ···················· | ···················· | ···················· | ·NNNNNNNNNNNNNNNNNNN | NNNNNNNNNNNNNNNNNNNN | 1 |
| V | 5357 | hCoV-19/Thailand/Bangkok-CONI-0291/2020\|EPI_ISL_69 | ···················· | ···················· | ···················· | ···················· | ··NNNNN·NNNN········ | NNNNNNNNNNNNNNN····· | 1 |
| V | 5358 | hCoV-19/Thailand/Bangkok-CONI-0293/2020\|EPI_ISL_69 | ···················· | ···················· | ···················· | ···················· | ··NNNNNNNNNNNNNNNNNN | NNNNNNNNNNNNNNNNNNNN | 1 |
| V | 5360 | hCoV-19/Thailand/Bangkok-CONI-0295/2020\|EPI_ISL_69 | ···················· | ···················· | ···················· | ···················· | NNNNNNNNNNNNNNNNNNNN | NNNNNNNNNNNNNNNNNNNN | 1 |
| V | 5361 | hCoV-19/Thailand/Bangkok-CONI-0297/2020\|EPI_ISL_69 | ···················· | ···················· | ···················· | ···················· | NNNNNNNNNNNNNNNNNNNN | NNNNNNNNNNNNNNNNNNNN | 1 |
| V | 5362 | hCoV-19/Thailand/Bangkok-CONI-0298/2020\|EPI_ISL_69 | ···················· | ···················· | ···················· | ···················· | ··NNNNNNNNNNNNNNNNNN | NNNNNNNNNNNNNNNNNNNN | 1 |
| V | 5364 | hCoV-19/Thailand/Bangkok-CONI-0300/2020\|EPI_ISL_69 | ···················· | ···················· | ···················· | ···················· | ··NNNNNNNNNNNNNNNNNN | ···················· | 1 |
| V | 5365 | hCoV-19/Thailand/Bangkok-CONI-0301/2020\|EPI_ISL_69 | ···················· | ···················· | ···················· | ···················· | ·NNNNNNNNNNNNNNNNNNN | NNNNNNNNNNNNNNNNNNNN | 1 |
| V | 5366 | hCoV-19/Thailand/Bangkok-CONI-0302/2020\|EPI_ISL_69 | ···················· | ···················· | ···················· | ···················· | ·NNNNNNNNNNNNNNNNNNN | NNNNNNNNNNNNNNNNNNNN | 1 |
| V | 5367 | hCoV-19/Thailand/Bangkok-CONI-0303/2020\|EPI_ISL_69 | ···················· | ···················· | ···················· | ···················· | ······NNNNNNNNNNNNNN | NNNNNNNNNNNNNNNNNNNN | 1 |
| V | 5368 | hCoV-19/Thailand/Bangkok-CONI-0304/2020\|EPI_ISL_69 | ···················· | ···················· | ···················· | ···················· | ··NNNNNNNNNNNNNNNNNN | ···················· | 1 |
| V | 5369 | hCoV-19/Thailand/Bangkok-CONI-0307/2020\|EPI_ISL_69 | ···················· | ···················· | ···················· | ···················· | ··NNNNNNNNNNNNNNNNNN | NNNNNNNNNNNNNNNNNNNN | 1 |
| V | 5370 | hCoV-19/Thailand/Bangkok-CONI-0308/2020\|EPI_ISL_69 | ···················· | ···················· | ···················· | ···················· | ··NNNNNNNNNNNNNNNNNN | NNNNNNNNNNNNNNNN···· | 1 |

| | | | | | | | | | |
|---|---|---|---|---|---|---|---|---|---|
| V | 5371 | hCoV-19/Thailand/Bangkok-CONI-0309/2020\|EPI_ISL_69 | ···················· | ···················· | ···················· | ···················· | ·NNNNNNNNNNNNNNNNNNN | NNNNNNNNNNNNNNNNNNNN | 1 |
| V | 5372 | hCoV-19/Thailand/Bangkok-CONI-0310/2020\|EPI_ISL_69 | ···················· | ···················· | ···················· | ···················· | ··NNNNNNNNNNNNNNNNNN | NNNNNNNNNNNNNNNNNNNN | 1 |
| V | 5373 | hCoV-19/Thailand/Bangkok-CONI-0311/2020\|EPI_ISL_69 | ···················· | ···················· | ···················· | ···················· | ··NNNNNNNNNNNNNNNNNN | NNNNNNNNNNNNNNNNNNNN | 1 |
| V | 5374 | hCoV-19/Thailand/Bangkok-CONI-0312/2020\|EPI_ISL_69 | ···················· | ···················· | ···················· | ···················· | ·NNNNNNNNNNNNNNNNNNN | NNNNNNNNNNNNNNNNNNNN | 1 |
| V | 5375 | hCoV-19/Thailand/Bangkok-CONI-0313/2020\|EPI_ISL_69 | ···················· | ···················· | ···················· | ···················· | ·NNNNNNNNNNNNNNNNNNN | NNNNNNNNNNNNNNNNNNNN | 1 |
| V | 5376 | hCoV-19/Thailand/Bangkok-CONI-0315/2020\|EPI_ISL_69 | ···················· | ···················· | ···················· | ···················· | ·NNNNNNNNNNNNNNNNNNN | NNNNNNNNNNNNNNNNNNNN | 1 |
| V | 5732 | hCoV-19/Japan/PG-8244/2020\|EPI_ISL_687343\|2020-08 | ···················· | ···················· | ···················· | ···················· | ···················· | NNNNNNNNNNNNNNNNNNNN | 1 |
| V | 5838 | hCoV-19/Japan/PG-5569/2020\|EPI_ISL_688694\|2020-08 | ···················· | NNNNNNNNNNNNNNNNNNNN | ···················· | ···················· | ···················· | ···················· | 1 |
| V | 6071 | hCoV-19/Japan/PG-2810/2020\|EPI_ISL_690937\|2020-04 | ···················· | ···················· | ···················· | ······NN·NNNNNNN···· | ···················· | ···················· | 1 |
| V | 6122 | hCoV-19/Japan/PG-2873/2020\|EPI_ISL_690999\|2020-04 | ···················· | ···················· | ···················· | ···················· | ···················· | NNNNNNNNNNNNNNNNNNNN | 1 |
| V | 6779 | hCoV-19/Japan/PG-7339/2020\|EPI_ISL_688442\|2020-08 | ···················· | ···················· | ···················· | ···················· | ···················· | NNNNNNNNNNNNNNNNNNNN | 1 |
| V | 7234 | hCoV-19/Japan/PG-7552/2020\|EPI_ISL_686654\|2020-09 | ···················· | ···················· | ···················· | ···················· | ·······NNNNNNNNNNNNN | NNNNNNNNNNNNNNNNNNNN | 1 |
| V | 7298 | hCoV-19/Japan/PG-6889/2020\|EPI_ISL_687997\|2020 | ···················· | ···················· | ···················· | ···················· | ···················· | NNNNNNNNNNNNNN······ | 1 |
| V | 7316 | hCoV-19/Japan/PG-3014/2020\|EPI_ISL_691138\|2020-04 | ···················· | ···················· | ···················· | ···················· | ···················· | NNNNN··············N | 1 |
| V | 7326 | hCoV-19/Japan/PG-3024/2020\|EPI_ISL_691148\|2020-04 | ···················· | ···················· | ···················· | ···················· | ···················· | NNNNNNNNNNNNNNNNNNNN | 1 |
| V | 7568 | hCoV-19/Japan/PG-2985/2020\|EPI_ISL_691109\|2020-04 | ···················· | ···················· | ···················· | ···················· | NNNNNNNNNNNNNNNNNNNN | NNNNNNNNNNNNNNNNNNNN | 1 |
| V | 7816 | hCoV-19/Japan/PG-2885/2020\|EPI_ISL_691011\|2020-04 | ···················· | ···················· | ···················· | ···················· | ···················· | NNNNNNNNNN·········· | 1 |
| V | 7819 | hCoV-19/Japan/PG-2888/2020\|EPI_ISL_691014\|2020-05 | ···················· | ···················· | ···················· | ···················· | ···················· | NNNNNNNNNNNNNNNNNNNN | 1 |
| V | 7882 | hCoV-19/Japan/PG-4112/2020\|EPI_ISL_692352\|2020-07 | ···················· | ···················· | ···················· | NNNNNNNNNNNNNNNNNNNN | ··············NNNNNN | NNNNNNNNNNNNNNNNNNNN | 1 |
| V | 8141 | hCoV-19/Japan/PG-4139/2020\|EPI_ISL_692378\|2020-07 | ···················· | ···················· | ···················· | ···················· | ···················· | NNNNNNNNNNNNNNNNNNNN | 1 |
| V | 8163 | hCoV-19/Portugal/PT1595/2020\|EPI_ISL_693600\|2020-0 | ···················· | ···················· | ···················· | ···················· | NNNNNNNNNNNNNNNNNNNN | NNNNNNNNNNNNNNNNNNNN | 1 |
| V | 8164 | hCoV-19/Portugal/PT1597/2020\|EPI_ISL_693601\|2020-0 | ···················· | ···················· | ···················· | ···················· | NNNNNNNNNNNNNNNNNNNN | ··N·NN··NN··NNN·NNN· | 1 |
| V | 8165 | hCoV-19/Portugal/PT1598/2020\|EPI_ISL_693602\|2020-0 | ···················· | ···················· | ···················· | ···················· | ·NNNNNNNNNNN··N····N | ···················· | 1 |
| V | 8169 | hCoV-19/Portugal/PT1603/2020\|EPI_ISL_693606\|2020-0 | ···················· | ···················· | ···················· | ···················· | ·NNNNNNNNNNNNNNNNNNN | ···················· | 1 |
| V | 8170 | hCoV-19/Portugal/PT1604/2020\|EPI_ISL_693607\|2020-0 | ···················· | ···················· | ···················· | ···················· | NNNNNNNNNNNNNNNNNNNN | ···················· | 1 |
| V | 8171 | hCoV-19/Portugal/PT1605/2020\|EPI_ISL_693608\|2020-0 | ···················· | ··········A········· | ···················· | ···················· | NNNNNNNNNNNNNNNNNNNN | ···················· | 1 |
| V | 8173 | hCoV-19/Portugal/PT1607/2020\|EPI_ISL_693610\|2020-0 | ···················· | ···················· | ···················· | ···················· | NNNNNNNNNNN·N·N····· | ···················· | 1 |
| V | 8176 | hCoV-19/Portugal/PT1611/2020\|EPI_ISL_693613\|2020-0 | ···················· | ···················· | ···················· | ···················· | N·N·NNNN·····N······ | ···················· | 1 |
| V | 8182 | hCoV-19/Portugal/PT1618/2020\|EPI_ISL_693619\|2020-0 | ···················· | ···················· | ···················· | ···················· | NNNNNNNNNNNNNNNNNNNN | NNNNNNNNNNNNNNNNNNNN | 1 |
| V | 8189 | hCoV-19/Portugal/PT1625/2020\|EPI_ISL_693626\|2020-0 | ···················· | ···················· | ···················· | ···················· | NNNNNNNNNNNNNNNNNNNN | NNNNNNNNNNNNNNNNNNNN | 1 |
| V | 8233 | hCoV-19/Japan/PG-1538/2020\|EPI_ISL_685144\|2020-04 | ···················· | ···················· | ···················· | ···················· | ···················· | NNNN················ | 1 |
| V | 8239 | hCoV-19/Japan/PG-1544/2020\|EPI_ISL_685150\|2020-04 | ···················· | ···················· | ···················· | ···················· | ···················· | NNNNNNNNNNNNNNNNNNNN | 1 |
| V | 8245 | hCoV-19/Japan/PG-1550/2020\|EPI_ISL_685156\|2020-04 | ···················· | ···················· | ···················· | ···················· | ···················· | NNNNNNNNNNNNNNNNNNNN | 1 |
| V | 8281 | hCoV-19/Japan/PG-8920/2020\|EPI_ISL_686466\|2020-09 | ···················· | ···················· | ···················· | ···················· | ···················· | NNNNNNNNNNNNNNNNNNNN | 1 |
| V | 8359 | hCoV-19/Japan/PG-3987/2020\|EPI_ISL_692228\|2020-07 | ···················· | ·A·················· | ···················· | ···················· | ···················· | ·····NNNNNNNNNNNNNNN | 1 |
| V | 8462 | hCoV-19/Japan/PG-1459/2020\|EPI_ISL_685066\|2020-04 | ···················· | ···················· | ···················· | ···················· | ···················· | NNNNNNNNNNNNNNNNNNNN | 1 |
| V | 8484 | hCoV-19/Japan/PG-1481/2020\|EPI_ISL_685088\|2020-04 | ···················· | ···················· | ···················· | ···················· | NNNNNNNNNNNNNNNNNNNN | NNNNNNNNNNNNNNNNNNNN | 1 |
| V | 8668 | hCoV-19/Portugal/PT1550/2020\|EPI_ISL_693563\|2020-0 | ···················· | ·········A·········· | ···················· | ···················· | NNNNNNNNNNNNNNNNNNNN | NNNNNNNNNNNNNNNNNNNN | 1 |
| V | 8669 | hCoV-19/Portugal/PT1551/2020\|EPI_ISL_693564\|2020-0 | ···················· | ···················· | ···················· | ···················· | NNNNNNNNNNNNNNNNNNNN | ··N··········N···NNN | 1 |
| V | 8683 | hCoV-19/Portugal/PT1569/2020\|EPI_ISL_693578\|2020-0 | ···················· | ···················· | ···················· | ···················· | NNNNNNNNNNNNNNNNNNNN | NNNNNNNNNNNNNNNNNNNN | 1 |
| V | 8695 | hCoV-19/Portugal/PT1584/2020\|EPI_ISL_693590\|2020-0 | ···················· | ···················· | ···················· | ···················· | NNNNNNNNNNNNNNNNNNNN | NNNNNNNNNNNNNNNNNNNN | 1 |
| V | 8696 | hCoV-19/Portugal/PT1585/2020\|EPI_ISL_693591\|2020-0 | ···················· | ···················· | ···················· | ···················· | NNNNNNNNNNNNNNNNNNNN | NNNNNNNNNNNNNNNNNNNN | 1 |
| V | 8697 | hCoV-19/Portugal/PT1587/2020\|EPI_ISL_693592\|2020-0 | ···················· | ···················· | ···················· | ···················· | NNNNNNNNNNNNNNNNNNNN | ···················· | 1 |
| V | 8699 | hCoV-19/Portugal/PT1589/2020\|EPI_ISL_693594\|2020-0 | ···················· | ···················· | ···················· | ···················· | ···················· | N·NNN·N····N········ | 1 |
| V | 8701 | hCoV-19/Portugal/PT1591/2020\|EPI_ISL_693596\|2020-0 | ···················· | ···················· | ···················· | ···················· | NNNNNNNNNNNNNNNNNNNN | NNNNNNNNNNNNNNNNNNNN | 1 |
| V | 8704 | hCoV-19/Portugal/PT1594/2020\|EPI_ISL_693599\|2020-0 | ···················· | ···················· | ···················· | ···················· | NNNNNNNNNNNNNNNNNNNN | NNNNNNNNNNNNNNNNNNNN | 1 |
| V | 8916 | hCoV-19/Japan/PG-3947/2020\|EPI_ISL_692196\|2020-07 | ···················· | ···················· | ···················· | ···················· | NNNNNNNNNNNNNNNNNNNN | NNNNNNNNNNNNNNNNNNNN | 1 |
| V | 9116 | hCoV-19/Japan/PG-3874/2020\|EPI_ISL_692125\|2020-06 | ···················· | ···················· | ···················· | ···················· | ···················· | NNNNNNNNNNNNNNNNNNNN | 1 |
| V | 9145 | hCoV-19/Japan/PG-3903/2020\|EPI_ISL_692154\|2020-07 | ···················· | ···················· | ···················· | ···················· | ···················· | NNNNNNNNNNNNNNNNNNNN | 1 |
| W | 56 | hCoV-19/South Africa/NHLS-UCT-GS-2090-KRISP/2020\|E | ···················· | ···················· | ···················· | ···················· | ··NNNNNNNNNNNNNNNNNN | NNNNNNNNNNNNNNNNNNNN | 1 |
| W | 57 | hCoV-19/South Africa/NHLS-UCT-GS-2101-KRISP/2020\|E | ···················· | ···················· | ···················· | ···················· | ··NNNNNNNNNNNNNNNNNN | NNNNNNNNNNNNNNNNNNNN | 1 |
| W | 58 | hCoV-19/South Africa/NHLS-UCT-GS-7001-KRISP/2020\|E | ···················· | ···················· | ···················· | ···················· | ··NNNNNNNNNNNNNNNNNN | NNNNNNNNNNNNNNNNNNNN | 1 |
| W | 59 | hCoV-19/South Africa/NHLS-UCT-GS-3558-KRISP/2020\|E | ···················· | ···················· | ···················· | ···················· | ··NNNNNNNNNNNNNNNNNN | NNNNNNNNNNNNNNNNNNNN | 1 |
| W | 60 | hCoV-19/South Africa/NHLS-UCT-GS-1392-KRISP/2020\|E | ···················· | ···················· | ···················· | ···················· | ·NNNNNNNNNNNNNNNNNNN | NNNNNNNNNNNNNNNNNNNN | 1 |
| W | 61 | hCoV-19/South Africa/NHLS-UCT-GS-1912-KRISP/2020\|E | ···················· | ···················· | ···················· | ···················· | ··NNNNNNNNNNNNNNNNNN | NNNNNNNNNNNNNNNNNNNN | 1 |
| W | 63 | hCoV-19/South Africa/NHLS-UCT-GS-7012-KRISP/2020\|E | ···················· | ···················· | ···················· | ···················· | ·NNNNNNNNNNNNNNNNNNN | NNNNNNNNNNNNNNNNNNNN | 1 |
| W | 65 | hCoV-19/South Africa/NHLS-UCT-GS-1851-KRISP/2020\|E | ···················· | ···················· | ···················· | ···················· | ··NNNNNNNNNNNNNNNNNN | NNNNNNNNNNNNNNNNNNNN | 1 |
| W | 67 | hCoV-19/South Africa/NHLS-UCT-GS-1454-KRISP/2020\|E | ···················· | ···················· | ···················· | ···················· | ··NNNNNNNNNNNNNNNNNN | NNNNNNNNNNNNNNNNNNNN | 1 |
| W | 68 | hCoV-19/South Africa/NHLS-UCT-GS-3467-KRISP/2020\|E | ···················· | ···················· | ···················· | ···················· | ··NNNNNNNNNNNNNNNNNN | NNNNNNNNNNNNNNNNNNNN | 1 |
| W | 69 | hCoV-19/South Africa/NHLS-UCT-GS-1396-KRISP/2020\|E | ···················· | ···················· | ···················· | ···················· | ······NNNNNNNNNNNNNN | NNNNNNNNNNNNNNNNNNNN | 1 |
| W | 70 | hCoV-19/South Africa/NHLS-UCT-GS-3556-KRISP/2020\|E | ···················· | ···················· | ···················· | ···················· | ··NNNNNNNNNNNNNNNNNN | ···················· | 1 |
| W | 71 | hCoV-19/South Africa/NHLS-UCT-GS-1395-KRISP/2020\|E | ···················· | ···················· | ···················· | ···················· | ··NNNNNNNNNNNNNNNNNN | NNNNNNNNNNNNNNNNNNNN | 1 |
| W | 72 | hCoV-19/South Africa/NHLS-UCT-GS-7003-KRISP/2020\|E | ···················· | ···················· | ···················· | ···················· | NNNNNNNNNNNNNNNNNNNN | NNNNNNNNNNNNNNNNNNNN | 1 |
| W | 73 | hCoV-19/South Africa/NHLS-UCT-GS-3553-KRISP/2020\|E | ···················· | ···················· | ···················· | ···················· | ·····NNNNNNNNNNNNNNN | NNNNNNNNNNNNNNNNNNNN | 1 |
| W | 74 | hCoV-19/South Africa/NHLS-UCT-GS-7019-KRISP/2020\|E | ···················· | ···················· | ···················· | ···················· | ·NNNNNNNNNNNNNNNNNNN | NNNNNNNNNNNNNNNNNNNN | 1 |
| W | 75 | hCoV-19/South Africa/NHLS-UCT-GS-3569-KRISP/2020\|E | ···················· | ···················· | ···················· | ···················· | ······NNNNNNNNNNNNNN | NNNNNNNNNNNNNNNNNNNN | 1 |
| W | 76 | hCoV-19/South Africa/NHLS-UCT-GS-7000-KRISP/2020\|E | ···················· | ···················· | ···················· | ···················· | ··NNNNNNNNNNNNNNNNNN | NNNNNNNNNNNNNNNNNNNN | 1 |
| W | 77 | hCoV-19/South Africa/NHLS-UCT-GS-7013-KRISP/2020\|E | ···················· | ···················· | ···················· | ···················· | ··NNNNNNNNNNNNNNNNNN | NNNNNNNNNNNNNNNNNNNN | 1 |
| W | 78 | hCoV-19/South Africa/NHLS-UCT-GS-7063-KRISP/2020\|E | ···················· | ···················· | ···················· | ···················· | ········NNNNNNNNNNNN | NNNNNNNNNNNNNNNNNNNN | 1 |
| W | 79 | hCoV-19/South Africa/NHLS-UCT-GS-7043-KRISP/2020\|E | ···················· | ···················· | ···················· | ···················· | ··NNNNNNNNNNNNNNNNNN | NNNNNNNNNNNNNNNNNNNN | 1 |
| W | 80 | hCoV-19/South Africa/NHLS-UCT-GS-7067-KRISP/2020\|E | ···················· | ···················· | ···················· | ···················· | ··NNNNNNNNNNNNNNNNNN | NNNNNNNNNNNNNNNNNNNN | 1 |
| W | 81 | hCoV-19/South Africa/NHLS-UCT-GS-7042-KRISP/2020\|E | ···················· | ···················· | ···················· | ···················· | ·NNNNNNNNNNNNNNNNNNN | NNNNNNNNNNNNNNNNNNNN | 1 |
| W | 82 | hCoV-19/South Africa/NHLS-UCT-GS-7038-KRISP/2020\|E | ···················· | ···················· | ···················· | ···················· | ··NNNNNNNNNNNNNNNNNN | NNNNNNNNNNNNNNNNNNNN | 1 |
| W | 186 | hCoV-19/England/NORW-F62AD/2020\|EPI_ISL_706362\|202 | ···················· | ···················· | ···················· | ···················· | NNNNNNNNNNNNNNNNNNNN | NNNNNNNNNNNNNNNNNNNN | 1 |
| W | 319 | hCoV-19/India/MH-ACTREC-558/2020\|EPI_ISL_700203\|20 | ···················· | ···················· | ···················· | ···················· | ···················· | NNNNNNNNNNNNNNNNNNNN | 1 |
| W | 325 | hCoV-19/India/MH-ACTREC-564/2020\|EPI_ISL_700209\|20 | ···················· | ···················· | ···················· | ···················· | ······NNNNNNNNNNNNNN | NN·················· | 1 |
| W | 331 | hCoV-19/India/MH-ACTREC-572/2020\|EPI_ISL_700216\|20 | ···················· | ···················· | ···················· | ···················· | ······NNNNNNNNNNNNNN | ···················· | 1 |
| W | 334 | hCoV-19/India/MH-ACTREC-576/2020\|EPI_ISL_700220\|20 | ···················· | ···················· | ···················· | ···················· | ······NNNNNNNNNNNNNN | NNNNNNNNNNNNNNNNNNNN | 1 |
| W | 340 | hCoV-19/India/MH-ACTREC-587/2020\|EPI_ISL_700226\|20 | ···················· | ···················· | ···················· | ···················· | ······NNNNNNNNNNNNNN | NNNNNNNNNNNNNNNNNNNN | 1 |
| W | 341 | hCoV-19/India/MH-ACTREC-588/2020\|EPI_ISL_700227\|20 | ···················· | ·A·················· | ···················· | ···················· | ···················· | ··············NNNNNN | 1 |

```
W   344 hCoV-19/India/MH-ACTREC-592/2020|EPI_ISL_700230|20   ····················  ····················  ····················  ····················  ·······NNNNNNNNNNNNN  ····················  1
W   349 hCoV-19/India/MH-ACTREC-597/2020|EPI_ISL_700235|20   ····················  ·A··················  ····················  ····················  ·······NNNNNNNNNNNNN  NNNNNNNNNNNNNNNNNNNN  1
W   354 hCoV-19/India/MH-ACTREC-603/2020|EPI_ISL_700240|20   ····················  ····················  ····················  ····················  ·······NNNNNNNNNNNNN  NNNNNNNNNNNNNNNNNNNN  1
W   356 hCoV-19/India/MH-ACTREC-605/2020|EPI_ISL_700242|20   ····················  ····················  ····················  ····················  ····················  NNNNNNNNNNNNNNNNNN··  1
W   357 hCoV-19/India/MH-ACTREC-606/2020|EPI_ISL_700243|20   ····················  ·A··················  ····················  ····················  ·······NNNNNNNNNNNNN  NNNNNNNNNNNNNNNNNNNN  1
W   367 hCoV-19/India/MH-ACTREC-618/2020|EPI_ISL_700253|20   ····················  ····················  ····················  ····················  ·······NNNNNNNNNNNNN  NNNNNNNNNNNNNNNNNNNN  1
W   377 hCoV-19/India/MH-ACTREC-628/2020|EPI_ISL_700263|20   ····················  ·····A··············  ····················  ····················  ········NNNNNNNNNNNN  ····················  1
W   381 hCoV-19/India/MH-ACTREC-634/2020|EPI_ISL_700267|20   ····················  ·A··················  ····················  ····················  ·······NNNNNNNNNNNNN  NNNNNNNNNNNNNNNNNNNN  1
W   386 hCoV-19/India/MH-ACTREC-639/2020|EPI_ISL_700272|20   ····················  ····················  ····················  ····················  ·······NNNNNNNNNNNNN  ····················  1
W   458 hCoV-19/England/NOTT-11F43E/2020|EPI_ISL_706432|20   ····················  ····················  ····················  ····················  NNNNNNNNNNNNNNNNNNNN  NNNNNNNNNNNNNNNNNNNN  1
W   459 hCoV-19/England/NOTT-11F50E/2020|EPI_ISL_706433|20   ····················  ····················  ····················  ····················  NNNNNNNNNNNNNNNNNNNN  NNNNNNNNNNNNNNNNNNNN  1
W   460 hCoV-19/England/NOTT-11F53B/2020|EPI_ISL_706434|20   ····················  ····················  ····················  ····················  NNNNNNNNNNNNNNNNNNNN  NNNNNNNNNNNNNNNNNNNN  1
W   464 hCoV-19/Wales/PHWC-48F014/2020|EPI_ISL_706438|2020   ····················  ····················  ····················  ····················  NNNNNNNNNNNNNNNNNNNN  ····················  1
W   465 hCoV-19/Wales/PHWC-48FFF9/2020|EPI_ISL_706439|2020   ····················  ····················  ····················  ····················  NNNNNNNNNNNNNNNNNNNN  NNNNNNNNNNNNNNNNNNNN  1
W   467 hCoV-19/Wales/PHWC-49003E/2020|EPI_ISL_706441|2020   ····················  ····················  ····················  ····················  NNNNNNNNNNNNNNNNNNNN  NNNNNNNNNNNNNNNNNNNN  1
W   468 hCoV-19/Wales/PHWC-49007A/2020|EPI_ISL_706442|2020   ····················  ····················  ····················  ····················  NNNNNNNNNNNNNNNNNNNN  NNNNNNNNNNNNNNNNNNNN  1
W   470 hCoV-19/Wales/PHWC-4900E3/2020|EPI_ISL_706444|2020   ····················  ····················  ····················  ····················  NNNNNNNNNNNNNNNNNNNN  NNNNNNNNNNNN········  1
W   473 hCoV-19/Wales/PHWC-49012C/2020|EPI_ISL_706447|2020   ····················  ····················  ····················  ····················  ····················  NNNNNNNNNNNNNNN·····  1
W   475 hCoV-19/Wales/PHWC-49014A/2020|EPI_ISL_706449|2020   ····················  ····················  ····················  ····················  NNNNNNNNNNNNNNNNNNNN  NNNNNNNNNNNNNNNNNNNN  1
W   479 hCoV-19/Thailand/Bangkok-CONI-0319/2020|EPI_ISL_70   ····················  ·A··················  ····················  ····················  ·NNNNNNNNNNNNNNNNNNN  NNNNNNNNNNNNNNNNNNNN  1
W   480 hCoV-19/Thailand/Bangkok-CONI-0321/2020|EPI_ISL_70   ····················  ·A··················  ····················  ····················  ·NNNNNNNNNNNNNNNNNNN  NNNNNNNNNNNNNNNNNNNN  1
W   481 hCoV-19/Thailand/Bangkok-CONI-0322/2020|EPI_ISL_70   ····················  ·A··················  ····················  ····················  ··NNNNNNNNNNNNNNNNNN  NNNNNNNNNNNNNNNNNNNN  1
W   482 hCoV-19/Thailand/Bangkok-CONI-0323/2020|EPI_ISL_70   ····················  ·A··················  ····················  ····················  NNNNNNNNNNNNNNNNNNNN  NNNNNNNNNNNNNNNNNNNN  1
W   696 hCoV-19/Wales/PHWC-4901C2/2020|EPI_ISL_706455|2020   ····················  ····················  ····················  ····················  NNNNNNNNNNNNNNNNNNNN  NNNNNNNNNNNNNNNNNNNN  1
W   704 hCoV-19/Wales/PHWC-490265/2020|EPI_ISL_706463|2020   ····················  ····················  ····················  ····················  NNNNNNNNNNNNNNNNNNNN  NNNNNNNNNNNNNNNNNNNN  1
W   705 hCoV-19/Wales/PHWC-490292/2020|EPI_ISL_706464|2020   ····················  ····················  ····················  ····················  NNNNNNNNNNNNNNNNNNNN  NNNNN·······NNNNNNNN  1
W   706 hCoV-19/Wales/PHWC-4902A1/2020|EPI_ISL_706465|2020   ····················  ····················  ····················  ····················  NNNNNNNNNNNNNNNNNNNN  NNNNNNNNNNNNNNNNNNNN  1
W   707 hCoV-19/Wales/PHWC-4902B0/2020|EPI_ISL_706466|2020   ····················  ····················  ····················  ····················  ····················  NNNNNNNNNNNNNNN··NNN  1
W   708 hCoV-19/Wales/PHWC-4902CF/2020|EPI_ISL_706467|2020   ····················  ····················  ····················  ····················  NNNNNNNNNNNNNNNNNNNN  NNNNNNNNNNNNNNNNNNNN  1
W   709 hCoV-19/Wales/PHWC-4902DE/2020|EPI_ISL_706468|2020   ····················  ····················  ····················  ····················  NNNNNNNNNNNNNNNNNNNN  NNNNNNNNNNNNNNNNNNNN  1
W   710 hCoV-19/Wales/PHWC-4902FC/2020|EPI_ISL_706469|2020   ····················  ····················  ····················  ····················  NNNNNNNNNNNNNNNNNNNN  NNNNN···············  1
W   711 hCoV-19/Wales/PHWC-490335/2020|EPI_ISL_706470|2020   ····················  ····················  ····················  ····················  NNNNNNNNNNNNNNNNNNNN  NNNNNNNNNNNNNNNNNNNN  1
W   712 hCoV-19/Wales/PHWC-490362/2020|EPI_ISL_706471|2020   ····················  ····················  ····················  ····················  NNNNNNNNNNNNNNNNNNNN  NNNNNNNNNNNNNNNNNNNN  1
W   713 hCoV-19/Wales/PHWC-490371/2020|EPI_ISL_706472|2020   ····················  ····················  ····················  ····················  NNNNNNNNNNNNNNNNNNNN  NNNNNNNNNNNNNNNNNNNN  1
W   715 hCoV-19/Wales/PHWC-4903AE/2020|EPI_ISL_706474|2020   ····················  ····················  ····················  ····················  NNNNNNNNNNNNNNNNNNNN  ····················  1
W   716 hCoV-19/Wales/PHWC-4903BD/2020|EPI_ISL_706475|2020   ····················  ····················  ····················  ····················  NNNNNNNNNNNNNNNNNNNN  NNNNNNNNNNNNNNNNNNNN  1
W   717 hCoV-19/Wales/PHWC-4903EA/2020|EPI_ISL_706476|2020   ····················  ····················  ····················  ····················  NNNNNNNNNNNNNNNNNNNN  NNNNNNNNNNNNNNNNNNNN  1
W   718 hCoV-19/Wales/PHWC-490405/2020|EPI_ISL_706477|2020   ····················  ····················  ····················  ····················  NNNNNNNNNNNNNNNNNNNN  NNNNNNNNNNNNNNNNNNNN  1
W   719 hCoV-19/Wales/PHWC-490414/2020|EPI_ISL_706478|2020   ····················  ····················  ····················  ····················  NNNNNNNNNNNNNNNNNNNN  ·NNNNNNNNNNNNNNNNNNN  1
W   720 hCoV-19/Wales/PHWC-490432/2020|EPI_ISL_706479|2020   ····················  ····················  ····················  ····················  NNNNNNNNNNNNNNNNNNNN  NNNNNNNNNNNNNNNNNNNN  1
W   721 hCoV-19/Wales/PHWC-490450/2020|EPI_ISL_706480|2020   ····················  ····················  ····················  ····················  NNNNNNNNNNNNNNNNNNNN  NNNNNNNNNNNNNNNNNNNN  1
W   722 hCoV-19/Wales/PHWC-49047E/2020|EPI_ISL_706481|2020   ····················  ····················  ····················  ····················  NNNNNNNNNNNNNNNNNNNN  NNNNNNNNNNNNNNNNNNNN  1
W   724 hCoV-19/Wales/PHWC-4904BA/2020|EPI_ISL_706483|2020   ····················  ····················  ····················  ····················  ······NNNNNN········  ····················  1
W   725 hCoV-19/Wales/PHWC-4904E7/2020|EPI_ISL_706484|2020   ····················  ····················  ····················  ····················  NNNNNNNNNNNNNNNNNNNN  NNNNNNNNNNNNNNNNNNNN  1
W   727 hCoV-19/Wales/PHWC-49053F/2020|EPI_ISL_706486|2020   ····················  ····················  ····················  ····················  NNNNNNNNNNNNNNNNNNNN  NNNNNNNNNNNNNNNNNNNN  1
W   729 hCoV-19/Wales/PHWC-49057B/2020|EPI_ISL_706488|2020   ····················  ····················  ····················  ····················  NNNNNNNNNNNNNNNNNNNN  ····················  1
W   731 hCoV-19/Wales/PHWC-4905B7/2020|EPI_ISL_706490|2020   ····················  ····················  ····················  ····················  NNNNNNNNNNNNNNNNNNNN  NNNNNNNNNNNNNNNNNNNN  1
W   734 hCoV-19/Wales/PHWC-4905F3/2020|EPI_ISL_706493|2020   ····················  ····················  ····················  ····················  NNNNNNNNNNNNNNNNNNNN  NNNNNNNNNNNNNNNNNNNN  1
W   735 hCoV-19/Wales/PHWC-49060F/2020|EPI_ISL_706494|2020   ····················  ····················  ····················  ····················  NNNNNNNNNNNNNNNNNNNN  NNNNNNNNNNNNNNNNNNNN  1
W   737 hCoV-19/Wales/PHWC-49063C/2020|EPI_ISL_706496|2020   ····················  ····················  ····················  ····················  NNNNNNNNNNNNNNNNNNNN  NNNNNNNNNNNNNNNNNNNN  1
W   738 hCoV-19/Wales/PHWC-49064B/2020|EPI_ISL_706497|2020   ····················  ····················  ····················  ····················  NNNNNNN·············  NNNNNNNNNNNNNNN·····  1
W   739 hCoV-19/Wales/PHWC-49065A/2020|EPI_ISL_706498|2020   ····················  ····················  ····················  ····················  NNNNNNNNNNNNNNNNNNNN  NNNNNNNNNN··········  1
W   816 hCoV-19/India/MH-ACTREC-693/2020|EPI_ISL_700318|20   ····················  ····················  ····················  ····················  ················NNNN  NNNNNNNNNNNNNNNNNNNN  1
W   822 hCoV-19/India/MH-ACTREC-699/2020|EPI_ISL_700324|20   ····················  ····················  ····················  ····················  ·······NNNNNNNNNNNNN  ····················  1
W   826 hCoV-19/Bangladesh/CHRF-0069/2020|EPI_ISL_700339|2   ····················  ····················  ····················  ····················  NNNNNNNNNNNNNNNNNNNN  NNNNNNNNNNNNNNNNNNNN  1
W   827 hCoV-19/Bangladesh/CHRF-0070/2020|EPI_ISL_700340|2   ····················  ····················  ····················  ····················  NNNNNNNNNNNNNNNNNNNN  NNNNNNNNNNNNNNNNNNNN  1
W   828 hCoV-19/Bangladesh/CHRF-0074/2020|EPI_ISL_700341|2   ····················  ····················  ····················  ····················  NNNNNNNNNNNNNNNNNNNN  NNNNNNNNNNNNNNNNNNNN  1
W   830 hCoV-19/Bangladesh/CHRF-0081/2020|EPI_ISL_700343|2   ····················  ····················  ····················  ····················  NNNNNNNNNNNNNNNNNNNN  NNNNNNNNNNNNNNNNNNNN  1
W   831 hCoV-19/Bangladesh/CHRF-0083/2020|EPI_ISL_700344|2   ····················  ····················  ····················  ····················  NNNNNNNNNNNNNNNNNNNN  ····················  1
W   832 hCoV-19/Bangladesh/CHRF-0084/2020|EPI_ISL_700345|2   ····················  ····················  ····················  ····················  NNNNNNNNNNNNNNNNNNNN  ····················  1
W   834 hCoV-19/Bangladesh/CHRF-0088/2020|EPI_ISL_700347|2   ····················  ····················  ····················  ····················  NNNNNN··············  ····················  1
W   925 hCoV-19/Wales/PHWC-4906C3/2020|EPI_ISL_706500|2020   ····················  ····················  ····················  ····················  NNNNNNNNNNNNNNNNNNNN  NNNNNNNNNNNNNNNNNNNN  1
W   927 hCoV-19/Wales/PHWC-4906F0/2020|EPI_ISL_706502|2020   ····················  ····················  ····················  ····················  NNNNNNNNNNNNNNNNNNNN  NNNNNNNNNNNNNNNNNNNN  1
W   928 hCoV-19/Wales/PHWC-49070C/2020|EPI_ISL_706503|2020   ····················  ····················  ····················  ····················  NNNNNNNNNNNNNNNNNNNN  ············NNNNNNNN  1
W   929 hCoV-19/Wales/PHWC-49071B/2020|EPI_ISL_706504|2020   ····················  ····················  ····················  ····················  ·············NNNNNN·  ····················  1
W   930 hCoV-19/Wales/PHWC-490739/2020|EPI_ISL_706505|2020   ····················  ····················  ····················  ····················  NNNNNNNNNNNNNNNNNNNN  NNNNNNNNNNNNNNNNNNNN  1
W   932 hCoV-19/Wales/PHWC-490757/2020|EPI_ISL_706507|2020   ····················  ····················  ····················  ····················  NNNNNNNNNNNNNNNNNNNN  NNNNNNNNNNNNNNNNNNNN  1
W   933 hCoV-19/Wales/PHWC-490766/2020|EPI_ISL_706508|2020   ····················  ····················  ····················  ····················  NNNNNNNNNNNNNNNNNNNN  NNNNNNNNNNNNNNNNNNNN  1
W   934 hCoV-19/Wales/PHWC-490784/2020|EPI_ISL_706509|2020   ····················  ····················  ····················  ····················  NNNNNNNNNNNNNNNNNNNN  NNNNNNNNNNNNNNNNNNNN  1
W   935 hCoV-19/Wales/PHWC-490793/2020|EPI_ISL_706510|2020   ····················  ····················  ····················  ····················  NNNNNNNNNNNNNNNNNNNN  NNNNNNNNNNNNNNNNNNNN  1
W   936 hCoV-19/Wales/PHWC-4907A2/2020|EPI_ISL_706511|2020   ····················  ····················  ····················  ····················  NNNNNNNNNNNNNNNNNNNN  NNNNNNNNNNNNNNN·····  1
W   937 hCoV-19/Wales/PHWC-4907B1/2020|EPI_ISL_706512|2020   ····················  ····················  ····················  ····················  ·····NNNNNNNN·······  ····················  1
W   940 hCoV-19/Wales/PHWC-490827/2020|EPI_ISL_706515|2020   ····················  ····················  ····················  ····················  NNNNNNNNNNNNNNNNNNNN  NNNNNNNNNNNNNNNNNNNN  1
W   941 hCoV-19/Wales/PHWC-490836/2020|EPI_ISL_706516|2020   ····················  ····················  ····················  ····················  NNNNNNNNNNNNNNNNNNNN  ···············NNNNN  1
W   943 hCoV-19/Wales/PHWC-490872/2020|EPI_ISL_706518|2020   ····················  ····················  ····················  ····················  NNNNNNNNNNNNNNNNNNNN  NNNNNNNNNNNNNNNNNNNN  1
W   944 hCoV-19/Wales/PHWC-490881/2020|EPI_ISL_706519|2020   ····················  ····················  ····················  ····················  ····················  NNNNNNNNNNNNNNN·····  1
W   945 hCoV-19/Wales/PHWC-490890/2020|EPI_ISL_706520|2020   ····················  ····················  ····················  ····················  NNNNNNNNNNNNNNNNNNNN  NNNNNNNNNNNNNNNNNNNN  1
W   946 hCoV-19/Wales/PHWC-4908AF/2020|EPI_ISL_706521|2020   ····················  ····················  ····················  ····················  NNNNNNNNNNNNNNNNNNNN  NNNNNNNNNNNNNNNNNNNN  1
W   948 hCoV-19/Wales/PHWC-4908DC/2020|EPI_ISL_706523|2020   ····················  ····················  ····················  ····················  ····················  NNNNN···············  1
W   949 hCoV-19/Wales/PHWC-4908EB/2020|EPI_ISL_706524|2020   ····················  ····················  ····················  ····················  NNNNNNNNNNNNNNNNNNNN  ····················  1
```

| | | | | | | | | |
|---|---|---|---|---|---|---|---|---|
| W | 951 hCoV-19/Wales/PHWC-490906/2020\|EPI_ISL_706526\|2020 | ···················· | ···················· | ···················· | ···················· | NNNNNNNNNNNNNNNNNNNN | NNNNNNNNNNNNNNNNNNNN | 1 |
| W | 952 hCoV-19/Wales/PHWC-490924/2020\|EPI_ISL_706527\|2020 | ···················· | ···················· | ···················· | ···················· | NNNNNNNNNNNNNNNNNNNN | NNNNNNNNNNNNNNNNNNNN | 1 |
| W | 953 hCoV-19/Wales/PHWC-490933/2020\|EPI_ISL_706528\|2020 | ···················· | ···················· | ···················· | ···················· | NNNNNNNNNNNNNNNNNNNN | NNNNNNNNNNNNNNNNNNNN | 1 |
| W | 954 hCoV-19/Wales/PHWC-49097F/2020\|EPI_ISL_706529\|2020 | ···················· | ···················· | ···················· | ···················· | ···················· | NNNNNNNNNNNNNNNNNNNN | 1 |
| W | 955 hCoV-19/Wales/PHWC-490C2B/2020\|EPI_ISL_706550\|2020 | ···················· | ···················· | ···················· | ···················· | ···················· | NNNNN·········N····· | 1 |
| W | 957 hCoV-19/Wales/PHWC-490C49/2020\|EPI_ISL_706552\|2020 | ···················· | ···················· | ···················· | ···················· | NNNNNNNNNNNNNNNNNNNN | NNNNN··············· | 1 |
| W | 959 hCoV-19/Wales/PHWC-490C67/2020\|EPI_ISL_706554\|2020 | ···················· | ···················· | ···················· | ···················· | NNNNNNNNNNNNNNNNNNNN | NNNNNNNNNNNNNNNNNNNN | 1 |
| W | 961 hCoV-19/Wales/PHWC-490C94/2020\|EPI_ISL_706556\|2020 | ···················· | ···················· | ···················· | ···················· | NNNNNNNNNNNNNNNNNNNN | NNNNN·······NNNNNNNN | 1 |
| W | 962 hCoV-19/Wales/PHWC-490CA3/2020\|EPI_ISL_706557\|2020 | ···················· | ···················· | ···················· | ···················· | NNNNNNNNNNNNNNNNNNNN | NNNNNNNNNNNNNNNNNNNN | 1 |
| W | 963 hCoV-19/Wales/PHWC-490CB2/2020\|EPI_ISL_706558\|2020 | ···················· | ···················· | ···················· | ···················· | NNNNNNNNNNNNNNNNNNNN | NNNNNNNNNNNNNNNNNNNN | 1 |
| W | 964 hCoV-19/Wales/PHWC-490CC1/2020\|EPI_ISL_706559\|2020 | ···················· | ···················· | ···················· | ···················· | NNNNNNNNNNNNNNNNNNNN | NNNNNNNNNNNNNNNNNNNN | 1 |
| W | 967 hCoV-19/New Zealand/20CV0639/2020\|EPI_ISL_707803\|2 | ···················· | ·A·················· | ···················· | ···················· | NNNNNNNNNNNNNNNNNNNN | NNNNNNNNNNNNNNNNNNNN | 1 |
| W | 969 hCoV-19/New Zealand/20CV0641/2020\|EPI_ISL_707805\|2 | ···················· | ···················· | ···················· | ···················· | NNNNNNNNNNNNNNNNNNNN | NNNNNNNNNNNNNNNNNNNN | 1 |
| W | 978 hCoV-19/USA/MI-UM-10036764267/2020\|EPI_ISL_707819\| | ···················· | ···················· | ···················· | ···················· | NNNNNNNNNNNNNNNNNNNN | ···················· | 1 |
| W | 980 hCoV-19/USA/MI-UM-10036795489/2020\|EPI_ISL_707821\| | ···················· | ···················· | ···················· | ···················· | NNNNNNNNNNNNNNNNNNNN | ···················· | 1 |
| W | 989 hCoV-19/USA/MI-UM-10036777156/2020\|EPI_ISL_707830\| | ···················· | ···················· | ···················· | ···················· | NNNNNNNNNNNNNNNNNNNN | ···················· | 1 |
| W | 990 hCoV-19/USA/MI-UM-10036799140/2020\|EPI_ISL_707831\| | ···················· | ···················· | ···················· | ···················· | ··NNNNNNNNNNNNNNNNNN | ···················· | 1 |
| W | 1005 hCoV-19/USA/MI-UM-10036802363/2020\|EPI_ISL_707846\| | ···················· | ·A·················· | ···················· | ···················· | NNNNNNNNNNNNNNNNNNNN | NNNNNNNNNNNNNNNNNNNN | 1 |
| W | 1011 hCoV-19/USA/MI-UM-10036803122/2020\|EPI_ISL_707852\| | ···················· | ·A·················· | ···················· | ···················· | NNNNNNNNNNNNNNNNNNNN | ···················· | 1 |
| W | 1012 hCoV-19/USA/MI-UM-10036799803/2020\|EPI_ISL_707853\| | ···················· | ···················· | ···················· | ···················· | NNNNNNNNNNNNNNNNNNNN | ···················· | 1 |
| W | 1013 hCoV-19/USA/MI-UM-10036801999/2020\|EPI_ISL_707854\| | ···················· | ···················· | ···················· | ···················· | NNNNNNNNNNNNNNNNNNNN | ···················· | 1 |
| W | 1015 hCoV-19/USA/MI-UM-10036797730/2020\|EPI_ISL_707856\| | ···················· | ···················· | ···················· | ···················· | NNNNNNNNNNNNNNNNNNNN | ···················· | 1 |
| W | 1020 hCoV-19/USA/MI-UM-10036801511/2020\|EPI_ISL_707881\| | ···················· | ···················· | ···················· | ···················· | ··········NNNNNNNNNN | ···················· | 1 |
| W | 1141 hCoV-19/France/ARA-SC144/2020\|EPI_ISL_700408\|2020- | ···················· | ·A·················· | ···················· | ···················· | NNNNNNNNNNNNN······· | ···················· | 1 |
| W | 1144 hCoV-19/South Africa/NHLS-UCT-GS-1330/2020\|EPI_ISL | ···················· | ···················· | ···················· | ···················· | ··NNNNNNNNNNNNNNNNNN | NNNNNNNNNNNNNNNNNNNN | 1 |
| W | 1145 hCoV-19/South Africa/NHLS-UCT-GS-1379/2020\|EPI_ISL | ···················· | ···················· | ···················· | ···················· | ··NNNNNNNNNNNNNNNNNN | NNNNNNNNNNNNNNNNNNNN | 1 |
| W | 1146 hCoV-19/South Africa/NHLS-UCT-GS-1049/2020\|EPI_ISL | ···················· | ···················· | ···················· | ···················· | ··NNNNNNNNNNNNNNNNNN | NNNNNNNNNNNNNNNNNNNN | 1 |
| W | 1147 hCoV-19/South Africa/NHLS-UCT-GS-1364/2020\|EPI_ISL | ···················· | ···················· | ···················· | ···················· | NNNNNNNNNNNNNNNNNNNN | NNNNNNNNNNNNNNNNNNNN | 1 |
| W | 1148 hCoV-19/South Africa/NHLS-UCT-GS-1332/2020\|EPI_ISL | ···················· | ···················· | ···················· | ···················· | ··NNNNNNNNNNNNNNNNNN | NNNNNNNNNNNNNNNNNNNN | 1 |
| W | 1194 hCoV-19/Wales/PHWC-490A21/2020\|EPI_ISL_706530\|2020 | ···················· | ···················· | ···················· | ···················· | ···················· | NNNNNNNNNNNNNNNNNNNN | 1 |
| W | 1195 hCoV-19/Wales/PHWC-490A30/2020\|EPI_ISL_706531\|2020 | ···················· | ···················· | ···················· | ···················· | NNNNNNNNNNNNNNNNNNNN | ···················· | 1 |
| W | 1196 hCoV-19/Wales/PHWC-490A4F/2020\|EPI_ISL_706532\|2020 | ···················· | ···················· | ···················· | ···················· | NNNNNNNNNNNNNNNNNNNN | NNNNNNNNNNNNNNNNNNNN | 1 |
| W | 1199 hCoV-19/Wales/PHWC-490AC7/2020\|EPI_ISL_706535\|2020 | ···················· | ···················· | ···················· | ···················· | NNNNNNNNN··········· | ···················· | 1 |
| W | 1202 hCoV-19/Wales/PHWC-490B1F/2020\|EPI_ISL_706538\|2020 | ···················· | ···················· | ···················· | ···················· | NNNNNNNNNNNNNNNNNNNN | NNNNNNNNNNNNNNNNNNNN | 1 |
| W | 1203 hCoV-19/Wales/PHWC-490B2E/2020\|EPI_ISL_706539\|2020 | ···················· | ·A·················· | ···················· | ···················· | NNNNNNNNNNNNNNNNNNNN | ··A················· | 1 |
| W | 1204 hCoV-19/Wales/PHWC-490B4C/2020\|EPI_ISL_706540\|2020 | ···················· | ···················· | ···················· | ···················· | NNNNNNNNNNNNNN······ | ···················· | 1 |
| W | 1206 hCoV-19/Wales/PHWC-490B79/2020\|EPI_ISL_706542\|2020 | ···················· | ···················· | ···················· | ···················· | NNNNNNNNNNNNNNNNNNNN | NNNNNNNNNNNNNNNNNNNN | 1 |
| W | 1207 hCoV-19/Wales/PHWC-490B88/2020\|EPI_ISL_706543\|2020 | ···················· | ···················· | ···················· | ···················· | NNNNNNNNNNNNNNNNNNNN | NNNNNNNNNNNNNNNNNNNN | 1 |
| W | 1209 hCoV-19/Wales/PHWC-490BD3/2020\|EPI_ISL_706545\|2020 | ···················· | ···················· | ···················· | ···················· | NNNNNNNNNNNNNNNNNNNN | ···················· | 1 |
| W | 1214 hCoV-19/Wales/PHWC-490CD0/2020\|EPI_ISL_706560\|2020 | ···················· | ···················· | ···················· | ···················· | NNNNNNNNNNNNNNNNNNNN | NNNNNNNNNNNNNNNNNNNN | 1 |
| W | 1215 hCoV-19/Wales/PHWC-490CEF/2020\|EPI_ISL_706561\|2020 | ···················· | ···················· | ···················· | ···················· | NNNNNNNNNNNNNNNNNNNN | NNNNNNNNNNNNNNNNNNNN | 1 |
| W | 1216 hCoV-19/Wales/PHWC-490D0A/2020\|EPI_ISL_706562\|2020 | ···················· | ···················· | ···················· | ···················· | NNNNNNNNNNNNNNNNNNNN | NNNNNNNNNNNNNNNNNNNN | 1 |
| W | 1217 hCoV-19/Wales/PHWC-490D19/2020\|EPI_ISL_706563\|2020 | ···················· | ···················· | ···················· | ···················· | NNNNNNNNNNNNNNNNNNNN | NNNNNNNNNNNNNNNNNNNN | 1 |
| W | 1219 hCoV-19/Wales/PHWC-490D37/2020\|EPI_ISL_706565\|2020 | ···················· | ···················· | ···················· | ···················· | NNNNNNNNNNNNNNNNNNNN | NNNNNNNNNNNNNNN····· | 1 |
| W | 1220 hCoV-19/Wales/PHWC-490D46/2020\|EPI_ISL_706566\|2020 | ···················· | ···················· | ···················· | ···················· | NNNNNNNNNNNNNNNNNNNN | NNNNNNNNNNNNNNNNNNNN | 1 |
| W | 1222 hCoV-19/Wales/PHWC-490D64/2020\|EPI_ISL_706568\|2020 | ···················· | ·····A·············· | ···················· | ···················· | NNNNNNNNNNNNNNNNNNNN | NNNNNNNNNNNNNNNNNNNN | 1 |
| W | 1223 hCoV-19/Wales/PHWC-490D73/2020\|EPI_ISL_706569\|2020 | ···················· | ···················· | ···················· | ···················· | NNNNNNNNNNNNNNNNNNNN | NNNNNNNNNNNNNNNNNNNN | 1 |
| W | 1224 hCoV-19/Wales/PHWC-490DA0/2020\|EPI_ISL_706570\|2020 | ···················· | ···················· | ···················· | ···················· | NNNNNNNNNNNNNNNNNNNN | NNNNNNNNNNNNNNNNNNNN | 1 |
| W | 1225 hCoV-19/Wales/PHWC-490DCE/2020\|EPI_ISL_706571\|2020 | ···················· | ···················· | ···················· | ···················· | NNNNNNNNNNNNNNNNNNNN | NNNNNNNNNNNNNNNNNNNN | 1 |
| W | 1227 hCoV-19/Wales/PHWC-490DFB/2020\|EPI_ISL_706573\|2020 | ···················· | ···················· | ···················· | ···················· | NNNNNNNNNNNNNNNNNNNN | NNNNNNNNNNNNNNNNNNNN | 1 |
| W | 1228 hCoV-19/Wales/PHWC-490E07/2020\|EPI_ISL_706574\|2020 | ···················· | ···················· | ···················· | ···················· | NNNNNNNNNNNNNNNNNNNN | NNNNNNNNNNNNNNNNNNNN | 1 |
| W | 1229 hCoV-19/Wales/PHWC-490E16/2020\|EPI_ISL_706575\|2020 | ···················· | ···················· | ···················· | ···················· | NNNNNNNNNNNNNNNNNNNN | NNNNNNNNNNNNNNNNNNNN | 1 |
| W | 1231 hCoV-19/Wales/PHWC-490E43/2020\|EPI_ISL_706577\|2020 | ···················· | ···················· | ···················· | ···················· | NNNNNNNNNNNNNNNNNNNN | NNNNNNNNNNNNNNNNN··· | 1 |
| W | 1232 hCoV-19/Wales/PHWC-490E52/2020\|EPI_ISL_706578\|2020 | ···················· | ···················· | ···················· | ···················· | NNNNNNNNNNNNNNNNNNNN | NNNNNNNNNNNNNNNNNNNN | 1 |
| W | 1241 hCoV-19/Wales/PHWC-490F22/2020\|EPI_ISL_706587\|2020 | ···················· | ···················· | ···················· | ···················· | NNNNNNNNNNNNNNNNNNNN | ···················· | 1 |
| W | 1255 hCoV-19/USA/MI-UM-10036786765/2020\|EPI_ISL_707861\| | ···················· | ···················· | ···················· | ···················· | NNNNNNNNNNNNNNNNNNNN | ···················· | 1 |
| W | 1256 hCoV-19/USA/MI-UM-10036774169/2020\|EPI_ISL_707862\| | ···················· | ···················· | ···················· | ···················· | NNNNNNNNNNNNNNNNNNNN | NNNNNNNNNNNNNNNNNNNN | 1 |
| W | 1257 hCoV-19/USA/MI-UM-10036789801/2020\|EPI_ISL_707863\| | ···················· | ···················· | ···················· | ···················· | NNNNNNNNNNNNNNNNNNNN | NNNNNNNNNNNNNNNNNNNN | 1 |
| W | 1258 hCoV-19/USA/MI-UM-10036785618/2020\|EPI_ISL_707864\| | ···················· | ·A·················· | ···················· | ···················· | NNNNNNNNNNNNNNNNNNNN | ···················· | 1 |
| W | 1262 hCoV-19/USA/MI-UM-10036782720/2020\|EPI_ISL_707868\| | ···················· | ···················· | ···················· | ···················· | NNNNNNNNNNNNNNNNNNNN | ···················· | 1 |
| W | 1269 hCoV-19/USA/MI-UM-10036788362/2020\|EPI_ISL_707875\| | ···················· | ···················· | ···················· | ···················· | NNNNNNNNNNNNNNNNNNNN | ···················· | 1 |
| W | 1320 hCoV-19/South Africa/NHLS-UCT-GS-1386/2020\|EPI_ISL | ···················· | ···················· | ···················· | ···················· | ···········NNNNNNNNN | NNNNNNNNNNNNNNNNNNNN | 1 |
| W | 1322 hCoV-19/South Africa/NHLS-UCT-GS-1369/2020\|EPI_ISL | ···················· | ···················· | ···················· | ···················· | NNNNNNNNNNNNNNNNNNNN | NNNNNNNNNNNNNNNNNNNN | 1 |
| W | 1323 hCoV-19/South Africa/NHLS-UCT-GS-1367/2020\|EPI_ISL | ···················· | ···················· | ···················· | ···················· | ··NNNNNNNNNNNNNNNNNN | NNNNNNNNNNNNNNNNNNNN | 1 |
| W | 1324 hCoV-19/South Africa/NHLS-UCT-GS-1100/2020\|EPI_ISL | ···················· | ···················· | ···················· | ···················· | NNNNNNNNNNNNNNNNNNNN | ···················· | 1 |
| W | 1326 hCoV-19/South Africa/NHLS-UCT-GS-1342/2020\|EPI_ISL | ···················· | ···················· | ···················· | ···················· | NNNNNNNNNNNNNNNNNNNN | NNNNNNNNNNNNNNNNNNNN | 1 |
| W | 1327 hCoV-19/South Africa/NHLS-UCT-GS-1374/2020\|EPI_ISL | ···················· | ···················· | ···················· | ···················· | ·NNNNNNNNNNNNNNNNNNN | NNNNNNNNNNNNNNNNNNNN | 1 |
| W | 1328 hCoV-19/South Africa/NHLS-UCT-GS-1025/2020\|EPI_ISL | ···················· | ···················· | ···················· | ···················· | NNNNNNNNNNNNNNNNNNNN | NNNNNNNNNNNNNNNNNNNN | 1 |
| W | 1329 hCoV-19/South Africa/NHLS-UCT-GS-1381/2020\|EPI_ISL | ···················· | ···················· | ···················· | ···················· | NNNNNNNNNNNNNNNNNNNN | ···················· | 1 |
| W | 1330 hCoV-19/South Africa/NHLS-UCT-GS-1382/2020\|EPI_ISL | ···················· | ···················· | ···················· | ···················· | ···············NNNNN | NNNNNNNNNNNNNNNNNNNN | 1 |
| W | 1332 hCoV-19/South Africa/NHLS-UCT-GS-1153/2020\|EPI_ISL | ···················· | ···················· | ···················· | ···················· | ·············NNNNNNN | NNNNNNNNNNNNNNNNNNNN | 1 |
| W | 1334 hCoV-19/South Africa/NHLS-UCT-GS-0808/2020\|EPI_ISL | ···················· | ···················· | ···················· | ···················· | ··NNNNNNNNNNNNNNNNNN | NNNNNNNNNNNNNNNNNNNN | 1 |
| W | 1335 hCoV-19/South Africa/NHLS-UCT-GS-1372/2020\|EPI_ISL | ···················· | ···················· | ···················· | ···················· | ··NNNNNNNNNNNNNNNNNN | NNNNNNNNNNNNNNNNNNNN | 1 |
| W | 1336 hCoV-19/South Africa/NHLS-UCT-GS-1384/2020\|EPI_ISL | ···················· | ···················· | ···················· | ···················· | ··NNNNNNNNNNNNNNNNNN | NNNNNNNNNNNNNNNNNNNN | 1 |
| W | 1337 hCoV-19/South Africa/NHLS-UCT-GS-1334/2020\|EPI_ISL | ···················· | ···················· | ···················· | ···················· | ··NNNNNNNNNNNNNNNNNN | NNNNNNNNNNNNNNNNNNNN | 1 |
| W | 1338 hCoV-19/South Africa/NHLS-UCT-GS-1366/2020\|EPI_ISL | ···················· | ···················· | ···················· | ···················· | NNNNNNNNNNNNNNNNNNNN | NNNNNNNNNNNNNNNNNNNN | 1 |
| W | 1343 hCoV-19/South Africa/NHLS-UCT-GS-2877/2020\|EPI_ISL | ···················· | ·A·················· | ···················· | ···················· | ·NNNNNNNNNNNNNNNNNNN | NNNNNNNNNNNNNNNNNNNN | 1 |
| W | 1344 hCoV-19/South Africa/NHLS-UCT-GS-1383/2020\|EPI_ISL | ···················· | ···················· | ···················· | ···················· | ·NNNNNNNNNNNNNNNNNNN | NNNNNNNNNNNNNNNNNNNN | 1 |
| W | 1346 hCoV-19/South Africa/NHLS-UCT-GS-1314/2020\|EPI_ISL | ···················· | ···················· | ···················· | ···················· | ··NNNNNNNNNNNNNNNNNN | NNNNNNNNNNNNNNNNNNNN | 1 |
| W | 1347 hCoV-19/South Africa/NHLS-UCT-GP-5157/2020\|EPI_ISL | ···················· | ·A·················· | ···················· | ···················· | NNNNNNNNNNNNNNNNNNNN | NNNNNNNNNNNNNNNNNNNN | 1 |

<table>
<tr><td>W</td><td>1348</td><td>hCoV-19/South Africa/NHLS-UCT-GS-0662/2020|EPI_ISL</td><td>····················</td><td>····················</td><td>····················</td><td>····················</td><td>···NNNNNNNNNNNNNNNNN</td><td>NNNNNNNNNNNNNNNNNNNN</td><td>1</td></tr>
<tr><td>W</td><td>1349</td><td>hCoV-19/South Africa/NHLS-UCT-GS-1269/2020|EPI_ISL</td><td>····················</td><td>····················</td><td>····················</td><td>····················</td><td>NNNNNNNNNNNNNNNNNNNN</td><td>NNNNNNNNNNNNNNNNNNNN</td><td>1</td></tr>
<tr><td>W</td><td>1352</td><td>hCoV-19/South Africa/NHLS-UCT-GS-1079/2020|EPI_ISL</td><td>····················</td><td>····················</td><td>····················</td><td>····················</td><td>·NNNNNNNNNNNNNNNNNNN</td><td>NNNNNNNNNNNNNNNNNNNN</td><td>1</td></tr>
<tr><td>W</td><td>1355</td><td>hCoV-19/South Africa/NHLS-UCT-GS-1148/2020|EPI_ISL</td><td>····················</td><td>····················</td><td>····················</td><td>····················</td><td>·NNNNNNNNNNNNNNNNNNN</td><td>NNNNNNNNNNNNNNNNNNNN</td><td>1</td></tr>
<tr><td>W</td><td>1357</td><td>hCoV-19/South Africa/NHLS-UCT-GS-0677/2020|EPI_ISL</td><td>····················</td><td>····················</td><td>····················</td><td>····················</td><td>··NNNNNNNNNNNNNNNNNN</td><td>NNNNNNNNNNNNNNNNNNNN</td><td>1</td></tr>
<tr><td>W</td><td>1358</td><td>hCoV-19/South Africa/NHLS-UCT-GS-1385/2020|EPI_ISL</td><td>····················</td><td>····················</td><td>····················</td><td>····················</td><td>··NNNNNNNNNNNNNNNNNN</td><td>NNNNNNNNNNNNNNNNNNNN</td><td>1</td></tr>
<tr><td>W</td><td>1363</td><td>hCoV-19/South Africa/NHLS-UCT-GS-1388/2020|EPI_ISL</td><td>····················</td><td>····················</td><td>····················</td><td>····················</td><td>NNNNNNNNNNNNNNNNNNNN</td><td>NNNNNNNNNNNNNNNNNNNN</td><td>1</td></tr>
<tr><td>W</td><td>1366</td><td>hCoV-19/South Africa/NHLS-UCT-GS-0633/2020|EPI_ISL</td><td>····················</td><td>····················</td><td>····················</td><td>····················</td><td>·NNNNNNNNNNNNNNNNNNN</td><td>NNNNNNNNNNNNNNNNNNNN</td><td>1</td></tr>
<tr><td>W</td><td>1369</td><td>hCoV-19/South Africa/NHLS-UCT-GS-2931/2020|EPI_ISL</td><td>····················</td><td>····················</td><td>····················</td><td>····················</td><td>NNNNNNNNNNNN········</td><td>····················</td><td>1</td></tr>
<tr><td>W</td><td>1370</td><td>hCoV-19/South Africa/NHLS-UCT-GS-0669/2020|EPI_ISL</td><td>····················</td><td>····················</td><td>····················</td><td>····················</td><td>··NNNNNNNNNNNNNNNNNN</td><td>NNNNNNNNNNNNNNNNNNNN</td><td>1</td></tr>
<tr><td>W</td><td>1371</td><td>hCoV-19/South Africa/NHLS-UCT-GS-0671/2020|EPI_ISL</td><td>····················</td><td>····················</td><td>····················</td><td>····················</td><td>··NNNNNNNNNNNNNNNNNN</td><td>NNNNNNNNNNNNNNNNNNNN</td><td>1</td></tr>
<tr><td>W</td><td>1373</td><td>hCoV-19/South Africa/NHLS-UCT-GS-1376/2020|EPI_ISL</td><td>····················</td><td>····················</td><td>····················</td><td>····················</td><td>··NNNNNNNNNNNNNNNNNN</td><td>NNNNNNNNNNNNNNNNNNNN</td><td>1</td></tr>
<tr><td>W</td><td>1375</td><td>hCoV-19/South Africa/NHLS-UCT-GS-1131/2020|EPI_ISL</td><td>····················</td><td>····················</td><td>····················</td><td>····················</td><td>········NNNNNNNNNNNN</td><td>NNNNNNNNNNNNNNNNNNNN</td><td>1</td></tr>
<tr><td>W</td><td>1378</td><td>hCoV-19/South Africa/NHLS-UCT-GS-1363/2020|EPI_ISL</td><td>····················</td><td>····················</td><td>····················</td><td>····················</td><td>··NNNNNNNNNNNNNNNNNN</td><td>NNNNNNNNNNNNNNNNNNNN</td><td>1</td></tr>
<tr><td>W</td><td>1380</td><td>hCoV-19/South Africa/NHLS-UCT-GS-0807/2020|EPI_ISL</td><td>····················</td><td>····················</td><td>····················</td><td>····················</td><td>············NNNNNNNN</td><td>····················</td><td>1</td></tr>
<tr><td>W</td><td>1384</td><td>hCoV-19/South Africa/NHLS-UCT-GS-1198/2020|EPI_ISL</td><td>····················</td><td>····················</td><td>····················</td><td>····················</td><td>·NNNNNNNNNNNNNNNNNNN</td><td>NNNNNNNNNNNNNNNNNNNN</td><td>1</td></tr>
<tr><td>W</td><td>1386</td><td>hCoV-19/South Africa/NHLS-UCT-GS-1030/2020|EPI_ISL</td><td>····················</td><td>····················</td><td>····················</td><td>····················</td><td>····················</td><td>NNNNNNNNNNNNNNNNNNNN</td><td>1</td></tr>
<tr><td>W</td><td>1427</td><td>hCoV-19/Wales/PHWC-49102E/2020|EPI_ISL_706600|2020</td><td>····················</td><td>····················</td><td>····················</td><td>····················</td><td>····················</td><td>NNNNNNNNNNNNNNN·····</td><td>1</td></tr>
<tr><td>W</td><td>1431</td><td>hCoV-19/Wales/PHWC-49106A/2020|EPI_ISL_706604|2020</td><td>····················</td><td>····················</td><td>····················</td><td>····················</td><td>NNNNNNNNNNNNNNNNNNNN</td><td>NNNNNNNNNNNNNNNNNNNN</td><td>1</td></tr>
<tr><td>W</td><td>1432</td><td>hCoV-19/Wales/PHWC-491088/2020|EPI_ISL_706605|2020</td><td>····················</td><td>····················</td><td>····················</td><td>····················</td><td>NNNNNNNNNNNNNNNNNNNN</td><td>····················</td><td>1</td></tr>
<tr><td>W</td><td>1434</td><td>hCoV-19/Wales/PHWC-4910A6/2020|EPI_ISL_706607|2020</td><td>····················</td><td>····················</td><td>····················</td><td>····················</td><td>NNNNNNNNNNNNNNNNNNNN</td><td>NNNNNNNNNNNNN·······</td><td>1</td></tr>
<tr><td>W</td><td>1435</td><td>hCoV-19/Wales/PHWC-4910B5/2020|EPI_ISL_706608|2020</td><td>····················</td><td>····················</td><td>····················</td><td>····················</td><td>····················</td><td>NNNNN···············</td><td>1</td></tr>
<tr><td>W</td><td>1437</td><td>hCoV-19/Wales/PHWC-4910D3/2020|EPI_ISL_706610|2020</td><td>····················</td><td>····················</td><td>····················</td><td>····················</td><td>NNNNNNNNNNNNNNNNNNNN</td><td>NNNNNNNNNNNNNNNNNNNN</td><td>1</td></tr>
<tr><td>W</td><td>1443</td><td>hCoV-19/Wales/PHWC-491149/2020|EPI_ISL_706616|2020</td><td>····················</td><td>····················</td><td>····················</td><td>····················</td><td>NNNNNNNNNNNNNNNNNNNN</td><td>····················</td><td>1</td></tr>
<tr><td>W</td><td>1448</td><td>hCoV-19/Wales/PHWC-4911A3/2020|EPI_ISL_706621|2020</td><td>····················</td><td>····················</td><td>····················</td><td>····················</td><td>NNNNNNNNNNNNNNNNNNNN</td><td>NNNNNNNNNNNNNNNNNNNN</td><td>1</td></tr>
<tr><td>W</td><td>1451</td><td>hCoV-19/Wales/PHWC-4911D0/2020|EPI_ISL_706624|2020</td><td>····················</td><td>····················</td><td>····················</td><td>····················</td><td>NNNNNNNNNNNNNNNNNNNN</td><td>NNNNNNNNNNNNNNNNNNNN</td><td>1</td></tr>
<tr><td>W</td><td>1461</td><td>hCoV-19/Wales/PHWC-491282/2020|EPI_ISL_706634|2020</td><td>····················</td><td>····················</td><td>····················</td><td>····················</td><td>NNNNNNNNNNNNNNNNNNNN</td><td>NNNNNNNNNNNNNNNNNNNN</td><td>1</td></tr>
<tr><td>W</td><td>1462</td><td>hCoV-19/Wales/PHWC-4912A0/2020|EPI_ISL_706635|2020</td><td>····················</td><td>····················</td><td>····················</td><td>····················</td><td>NNNNNNNNNNNNNNNNNNNN</td><td>····················</td><td>1</td></tr>
<tr><td>W</td><td>1469</td><td>hCoV-19/Wales/PHWC-491316/2020|EPI_ISL_706642|2020</td><td>····················</td><td>····················</td><td>····················</td><td>····················</td><td>NNNNNNNNNNNNNNNNNNNN</td><td>NNNNNNNNNNNNNNNNNNNN</td><td>1</td></tr>
<tr><td>W</td><td>1475</td><td>hCoV-19/Wales/PHWC-4913CB/2020|EPI_ISL_706648|2020</td><td>····················</td><td>····················</td><td>····················</td><td>····················</td><td>N···················</td><td>NNNNNNNNNNNNNNN·····</td><td>1</td></tr>
<tr><td>W</td><td>1477</td><td>hCoV-19/Wales/PHWC-4915F2/2020|EPI_ISL_706670|2020</td><td>····················</td><td>····················</td><td>····················</td><td>····················</td><td>NNNNNNNNNNNNNNNNNNNN</td><td>NNNNNNNNNNNNNNNNNNNN</td><td>1</td></tr>
<tr><td>W</td><td>1478</td><td>hCoV-19/Wales/PHWC-49161D/2020|EPI_ISL_706671|2020</td><td>····················</td><td>····················</td><td>····················</td><td>····················</td><td>NNNNNNNNNNNNNNNNNNNN</td><td>NNNNNNNNNNNNNNNNNNNN</td><td>1</td></tr>
<tr><td>W</td><td>1479</td><td>hCoV-19/Wales/PHWC-49162C/2020|EPI_ISL_706672|2020</td><td>····················</td><td>····················</td><td>····················</td><td>····················</td><td>NNNNNNNNNNNNNNNNNNNN</td><td>NNNNNNNNNNNNNNNNNNNN</td><td>1</td></tr>
<tr><td>W</td><td>1480</td><td>hCoV-19/Wales/PHWC-49163B/2020|EPI_ISL_706673|2020</td><td>····················</td><td>····················</td><td>····················</td><td>····················</td><td>NNNNNNNNNNNNNNNNNNNN</td><td>NNNNNNNNNNNNNNNNNNNN</td><td>1</td></tr>
<tr><td>W</td><td>1481</td><td>hCoV-19/Wales/PHWC-49164A/2020|EPI_ISL_706674|2020</td><td>····················</td><td>····················</td><td>····················</td><td>····················</td><td>NNNNNNNNNNNNNNNNNNNN</td><td>NNNNNNNNNNNNNNNNNNNN</td><td>1</td></tr>
<tr><td>W</td><td>1533</td><td>hCoV-19/Germany/SL-SU-10450116/2020|EPI_ISL_707970</td><td>····················</td><td>····················</td><td>····················</td><td>····················</td><td>····················</td><td>NNNNNNNNNNNNNNNNNNNN</td><td>1</td></tr>
<tr><td>W</td><td>1628</td><td>hCoV-19/South Africa/NHLS-UCT-GS-1312/2020|EPI_ISL</td><td>····················</td><td>····················</td><td>····················</td><td>····················</td><td>·NNNNNNNNNNNNNNNNNNN</td><td>NNNNNNNNNNNNNNNNNNNN</td><td>1</td></tr>
<tr><td>W</td><td>1630</td><td>hCoV-19/South Africa/NHLS-UCT-GS-1365/2020|EPI_ISL</td><td>····················</td><td>····················</td><td>····················</td><td>····················</td><td>·NNNNNNNNNNNNNNNNNNN</td><td>NNNNNNNNNNNNNNNNNNNN</td><td>1</td></tr>
<tr><td>W</td><td>1632</td><td>hCoV-19/South Africa/NHLS-UCT-GS-0676/2020|EPI_ISL</td><td>····················</td><td>····················</td><td>····················</td><td>····················</td><td>NNNNNNNNNNNNNNNNNNNN</td><td>NNNNNNNNNNNNNNNNNNNN</td><td>1</td></tr>
<tr><td>W</td><td>1642</td><td>hCoV-19/South Africa/NHLS-UCT-GS-1389/2020|EPI_ISL</td><td>····················</td><td>····················</td><td>····················</td><td>····················</td><td>NNNNNNNNNNNNNNNNNNNN</td><td>NNNNNNNNNNNNNNNNNNNN</td><td>1</td></tr>
<tr><td>W</td><td>1646</td><td>hCoV-19/South Africa/NHLS-UCT-GS-1380/2020|EPI_ISL</td><td>····················</td><td>····················</td><td>····················</td><td>····················</td><td>·NNNNNNNNNNNNNNNNNNN</td><td>NNNNNNNNNNNNNNNNNNNN</td><td>1</td></tr>
<tr><td>W</td><td>1739</td><td>hCoV-19/Wales/PHWC-4913F8/2020|EPI_ISL_706651|2020</td><td>····················</td><td>····················</td><td>····················</td><td>····················</td><td>NNNNNNNNNNNNNNNNNNNN</td><td>NNNNNNNNNNNNNNNNNN··</td><td>1</td></tr>
<tr><td>W</td><td>1740</td><td>hCoV-19/Wales/PHWC-491404/2020|EPI_ISL_706652|2020</td><td>····················</td><td>····················</td><td>····················</td><td>····················</td><td>NNNNNNNNNNNNNNNNNNNN</td><td>····················</td><td>1</td></tr>
<tr><td>W</td><td>1741</td><td>hCoV-19/Wales/PHWC-491413/2020|EPI_ISL_706653|2020</td><td>····················</td><td>····················</td><td>····················</td><td>····················</td><td>NNNNNN··············</td><td>NNNNNNNNNNNNNNNNNNNN</td><td>1</td></tr>
<tr><td>W</td><td>1744</td><td>hCoV-19/Wales/PHWC-49147D/2020|EPI_ISL_706656|2020</td><td>····················</td><td>····················</td><td>····················</td><td>····················</td><td>NNNNNNNNNNNNNNNNNNNN</td><td>NNNNNNNNNNNNNNNNNNNN</td><td>1</td></tr>
<tr><td>W</td><td>1747</td><td>hCoV-19/Wales/PHWC-4914AA/2020|EPI_ISL_706659|2020</td><td>····················</td><td>····················</td><td>····················</td><td>····················</td><td>····················</td><td>·····NNNNNN·········</td><td>1</td></tr>
<tr><td>W</td><td>1751</td><td>hCoV-19/Wales/PHWC-491510/2020|EPI_ISL_706663|2020</td><td>····················</td><td>····················</td><td>····················</td><td>····················</td><td>NNNNNNNNNNNNNNNNNNNN</td><td>NNNNNNNNNNNNNNNNNNNN</td><td>1</td></tr>
<tr><td>W</td><td>1752</td><td>hCoV-19/Wales/PHWC-49152F/2020|EPI_ISL_706664|2020</td><td>····················</td><td>····················</td><td>····················</td><td>····················</td><td>NNNNNNNNNNNNNNNNNNNN</td><td>NNNNNNN·············</td><td>1</td></tr>
<tr><td>W</td><td>1753</td><td>hCoV-19/Wales/PHWC-49153E/2020|EPI_ISL_706665|2020</td><td>····················</td><td>····················</td><td>····················</td><td>····················</td><td>NNNNNNNNNNNNNNNNNNNN</td><td>NNNNNNNNNNNNNNNNNNNN</td><td>1</td></tr>
<tr><td>W</td><td>1754</td><td>hCoV-19/Wales/PHWC-49156B/2020|EPI_ISL_706666|2020</td><td>····················</td><td>····················</td><td>····················</td><td>····················</td><td>NNNNNNNNNNNNNNNNNNNN</td><td>NNNNNNNNNNNNNNNNNNNN</td><td>1</td></tr>
<tr><td>W</td><td>1755</td><td>hCoV-19/Wales/PHWC-49157A/2020|EPI_ISL_706667|2020</td><td>····················</td><td>····················</td><td>····················</td><td>····················</td><td>NNNNNNNNNNNNNNNNNNNN</td><td>NNNNNNNNNNNNNNNNNNNN</td><td>1</td></tr>
<tr><td>W</td><td>1756</td><td>hCoV-19/Wales/PHWC-4915A7/2020|EPI_ISL_706668|2020</td><td>····················</td><td>····················</td><td>····················</td><td>····················</td><td>NNNNNNNNNNNNNNNNNNNN</td><td>NNNNNNNNNNNNNNNNNNNN</td><td>1</td></tr>
<tr><td>W</td><td>1757</td><td>hCoV-19/Wales/PHWC-4915D4/2020|EPI_ISL_706669|2020</td><td>····················</td><td>····················</td><td>····················</td><td>····················</td><td>NNNNNNNNNNNNNNNNNNNN</td><td>NNNNNNNNNNNNNNNNNNNN</td><td>1</td></tr>
<tr><td>W</td><td>1759</td><td>hCoV-19/Wales/PHWC-4916B3/2020|EPI_ISL_706679|2020</td><td>····················</td><td>····················</td><td>····················</td><td>····················</td><td>NNNNNNNNNNNNNNNNNNNN</td><td>·········NNNNNN·····</td><td>1</td></tr>
<tr><td>W</td><td>1762</td><td>hCoV-19/Wales/PHWC-4916FF/2020|EPI_ISL_706682|2020</td><td>····················</td><td>····················</td><td>····················</td><td>····················</td><td>NNNNNNNNNNNNNNNNNNNN</td><td>NNNNNNNNNNNNNNNNNNNN</td><td>1</td></tr>
<tr><td>W</td><td>1765</td><td>hCoV-19/Wales/PHWC-491729/2020|EPI_ISL_706685|2020</td><td>····················</td><td>····················</td><td>····················</td><td>····················</td><td>NNNNNNNNNNNNNNNNNNNN</td><td>·················NNN</td><td>1</td></tr>
<tr><td>W</td><td>1767</td><td>hCoV-19/Wales/PHWC-491747/2020|EPI_ISL_706687|2020</td><td>····················</td><td>····················</td><td>····················</td><td>····················</td><td>NNNNNNNNNNNNNNNNNNNN</td><td>····················</td><td>1</td></tr>
<tr><td>W</td><td>1768</td><td>hCoV-19/Wales/PHWC-491756/2020|EPI_ISL_706688|2020</td><td>····················</td><td>····················</td><td>····················</td><td>····················</td><td>NNNNNNNNNNNNNNNNNNNN</td><td>····················</td><td>1</td></tr>
<tr><td>W</td><td>1773</td><td>hCoV-19/Wales/PHWC-4917A1/2020|EPI_ISL_706693|2020</td><td>····················</td><td>····················</td><td>····················</td><td>····················</td><td>····················</td><td>NNNNNNNNNNNNNNN·NNNN</td><td>1</td></tr>
<tr><td>W</td><td>1777</td><td>hCoV-19/Wales/PHWC-491817/2020|EPI_ISL_706697|2020</td><td>····················</td><td>····················</td><td>····················</td><td>····················</td><td>····················</td><td>NNNNN···············</td><td>1</td></tr>
<tr><td>W</td><td>1788</td><td>hCoV-19/Germany/SL-SU-10451535/2020|EPI_ISL_707994</td><td>····················</td><td>····················</td><td>····················</td><td>····················</td><td>····················</td><td>NNNNNNNNNNNNNNNNNNNN</td><td>1</td></tr>
<tr><td>W</td><td>1847</td><td>hCoV-19/South Africa/NHLS-UCT-GS-1107/2020|EPI_ISL</td><td>····················</td><td>····················</td><td>····················</td><td>····················</td><td>·····NNNNNNNNNNNNNNN</td><td>NNNNNNNNNNNNNNNNNNNN</td><td>1</td></tr>
<tr><td>W</td><td>1848</td><td>hCoV-19/South Africa/NHLS-UCT-GS-1377/2020|EPI_ISL</td><td>····················</td><td>····················</td><td>····················</td><td>····················</td><td>··NNNNNNNNNNNNNNNNNN</td><td>NNNNNNNNNNNNNNNNNNNN</td><td>1</td></tr>
<tr><td>W</td><td>1856</td><td>hCoV-19/South Africa/NHLS-UCT-GS-1344/2020|EPI_ISL</td><td>····················</td><td>····················</td><td>····················</td><td>····················</td><td>NNNNNNNNNNNNNNNNNNNN</td><td>NNNNNNNNNNNNNNNNNNNN</td><td>1</td></tr>
<tr><td>W</td><td>1858</td><td>hCoV-19/South Africa/NHLS-UCT-GS-1349/2020|EPI_ISL</td><td>····················</td><td>····················</td><td>····················</td><td>····················</td><td>··NNNNNNNNNNNNNNNNNN</td><td>NNNNNNNNNNNNNNNNNNNN</td><td>1</td></tr>
<tr><td>W</td><td>1862</td><td>hCoV-19/South Africa/NHLS-UCT-GS-0684/2020|EPI_ISL</td><td>····················</td><td>····················</td><td>····················</td><td>····················</td><td>··NNNNNNNNNNNNNNNNNN</td><td>NNNNNNNNNNNNNNNNNNNN</td><td>1</td></tr>
<tr><td>W</td><td>1866</td><td>hCoV-19/South Africa/NHLS-UCT-GS-1047/2020|EPI_ISL</td><td>····················</td><td>····················</td><td>····················</td><td>····················</td><td>·NNNNNNNNNNNNNNNNNNN</td><td>NNNNNNNNNNNNNNNNNNNN</td><td>1</td></tr>
<tr><td>W</td><td>1867</td><td>hCoV-19/South Africa/NHLS-UCT-GS-1071/2020|EPI_ISL</td><td>····················</td><td>····················</td><td>····················</td><td>····················</td><td>··NNNNNNNNNNNNNNNNNN</td><td>NNNNNNNNNNNNNNNNNNNN</td><td>1</td></tr>
<tr><td>W</td><td>1868</td><td>hCoV-19/South Africa/NHLS-UCT-GS-1373/2020|EPI_ISL</td><td>····················</td><td>····················</td><td>····················</td><td>····················</td><td>··NNNNNNNNNNNNNNNNNN</td><td>NNNNNNNNNNNNNNNNNNNN</td><td>1</td></tr>
<tr><td>W</td><td>1869</td><td>hCoV-19/South Africa/NHLS-UCT-GS-1378/2020|EPI_ISL</td><td>····················</td><td>····················</td><td>····················</td><td>····················</td><td>·NNNNNNNNNNNNNNNNNNN</td><td>NNNNNNNNNNNNNNNNNNNN</td><td>1</td></tr>
<tr><td>W</td><td>1877</td><td>hCoV-19/South Africa/NHLS-UCT-GS-1362/2020|EPI_ISL</td><td>····················</td><td>····················</td><td>····················</td><td>····················</td><td>·NNNNNNNNNNNNNNNNNNN</td><td>NNNNNNNNNNNNNNNNNNNN</td><td>1</td></tr>
<tr><td>W</td><td>1878</td><td>hCoV-19/South Africa/NHLS-UCT-GS-0812/2020|EPI_ISL</td><td>····················</td><td>····················</td><td>····················</td><td>····················</td><td>NNNNNNNNNNNNNNNNNNNN</td><td>NNNNNNNNNNNNNNNNNNNN</td><td>1</td></tr>
<tr><td>W</td><td>1880</td><td>hCoV-19/South Africa/NHLS-UCT-GS-1449/2020|EPI_ISL</td><td>····················</td><td>·A··················</td><td>····················</td><td>····················</td><td>NNNNNNNNNNN·········</td><td>NNNNNNNNNNNNNNN·····</td><td>1</td></tr>
<tr><td>W</td><td>1881</td><td>hCoV-19/South Africa/NHLS-UCT-GS-1145/2020|EPI_ISL</td><td>····················</td><td>····················</td><td>····················</td><td>····················</td><td>··NNNNNNNNNNNNNNNNNN</td><td>NNNNNNNNNNNNNNNNNNNN</td><td>1</td></tr>
<tr><td>W</td><td>1890</td><td>hCoV-19/South Africa/NHLS-UCT-GS-1042/2020|EPI_ISL</td><td>····················</td><td>····················</td><td>····················</td><td>····················</td><td>··NNNNNNNNNNNNNNNNNN</td><td>NNNNNNNNNNNNNNNNNNNN</td><td>1</td></tr>
<tr><td>W</td><td>1906</td><td>hCoV-19/South Africa/NHLS-UCT-GS-1326/2020|EPI_ISL</td><td>····················</td><td>····················</td><td>····················</td><td>····················</td><td>··NNNNNNNNNNNNNNNNNN</td><td>NNNNNNNNNNNNNNNNNNNN</td><td>1</td></tr>
<tr><td>W</td><td>1908</td><td>hCoV-19/South Africa/NHLS-UCT-GS-0659/2020|EPI_ISL</td><td>····················</td><td>····················</td><td>····················</td><td>····················</td><td>··NNNNNNNNNNNNNNNNNN</td><td>NNNNNNNNNNNNNNNNNNNN</td><td>1</td></tr>
<tr><td>W</td><td>1914</td><td>hCoV-19/South Africa/NHLS-UCT-GS-0666/2020|EPI_ISL</td><td>····················</td><td>····················</td><td>····················</td><td>····················</td><td>··NNNNNNNNNNNNNNNNNN</td><td>NNNNNNNNNNNNNNNNNNNN</td><td>1</td></tr>
<tr><td>W</td><td>1916</td><td>hCoV-19/South Africa/NHLS-UCT-GS-1390/2020|EPI_ISL</td><td>····················</td><td>····················</td><td>····················</td><td>····················</td><td>NNNNNNNNNNNNNNNNNNNN</td><td>NNNNNNNNNNNNNNNNNNNN</td><td>1</td></tr>
<tr><td>W</td><td>1917</td><td>hCoV-19/South Africa/NHLS-UCT-GS-1028/2020|EPI_ISL</td><td>····················</td><td>····················</td><td>····················</td><td>····················</td><td>·NNNNNNNNNNNNNNNNNNN</td><td>NNNNNNNNNNNNNNNNNNNN</td><td>1</td></tr>
</table>

| | | | | | | | | |
|---|---|---|---|---|---|---|---|---|
| W | 1985 hCoV-19/Wales/PHWC-490344/2020\|EPI_ISL_705433\|2020 | ···················· | ···················· | ···················· | ···················· | NNNNNNNNNNNNNNNNNNNN | NNNNNNNNNNNNNNNNNNNN | 1 |
| W | 1996 hCoV-19/Wales/PHWC-4918AE/2020\|EPI_ISL_706704\|2020 | ···················· | ···················· | ···················· | ···················· | NNNNNNNNNNNNNNNNNNNN | NNNNNNNNNNNNNNNNNNNN | 1 |
| W | 1998 hCoV-19/Wales/PHWC-4918CC/2020\|EPI_ISL_706706\|2020 | ···················· | ···················· | ···················· | ···················· | NNNNNNNNNNNNNNNNNNNN | NNNNNNNNNNNNNNNNNNNN | 1 |
| W | 1999 hCoV-19/Wales/PHWC-4918DB/2020\|EPI_ISL_706707\|2020 | ···················· | ···················· | ···················· | ···················· | NNNNNNNNNNNNNNNNNNNN | NNNNNNNNNNNNNNNNNNNN | 1 |
| W | 2000 hCoV-19/Wales/PHWC-4918EA/2020\|EPI_ISL_706708\|2020 | ···················· | ···················· | ···················· | ···················· | NNNNNNNNNNNNNNNNNNNN | NNNNNNNNNNNNNNNNNNNN | 1 |
| W | 2001 hCoV-19/Wales/PHWC-4918F9/2020\|EPI_ISL_706709\|2020 | ···················· | ···················· | ···················· | ···················· | NNNNNNNNNNNNNNNNNNNN | NNNNNNNNNNNNNNNNNNNN | 1 |
| W | 2005 hCoV-19/Wales/PHWC-491932/2020\|EPI_ISL_706713\|2020 | ···················· | ···················· | ···················· | ···················· | NNNNNNNNNNNNNNNNNNNN | NNNNNNNNNNNNNNNNN··· | 1 |
| W | 2009 hCoV-19/Wales/PHWC-49197E/2020\|EPI_ISL_706717\|2020 | ···················· | ···················· | ···················· | ···················· | ···················· | NNNNNNNNNNNNNNN····· | 1 |
| W | 2010 hCoV-19/Wales/PHWC-49198D/2020\|EPI_ISL_706718\|2020 | ···················· | ···················· | ···················· | ···················· | NNNNNNNNNNNNNNNNNNNN | NNNNNNNNNNNNNNNNNNNN | 1 |
| W | 2012 hCoV-19/Wales/PHWC-4919AB/2020\|EPI_ISL_706720\|2020 | ···················· | ···················· | ···················· | ···················· | NNNNNNNNNNNNNNNNNNNN | NNNNNNNNNNNNNNNNNNNN | 1 |
| W | 2015 hCoV-19/Wales/PHWC-4919D8/2020\|EPI_ISL_706723\|2020 | ···················· | ···················· | ···················· | ···················· | NNNNNNNNNNNNNNNNNNNN | NNNNNNNNNNNNNNNNNNNN | 1 |
| W | 2016 hCoV-19/Wales/PHWC-4919E7/2020\|EPI_ISL_706724\|2020 | ···················· | ···················· | ···················· | ···················· | NNNNNNNNNNNNNN······ | ···················· | 1 |
| W | 2017 hCoV-19/Wales/PHWC-4919F6/2020\|EPI_ISL_706725\|2020 | ···················· | ···················· | ···················· | ···················· | NNNNNNNNNNNNNNNNNNNN | NNNNNNNNNNNNNNNNNNNN | 1 |
| W | 2019 hCoV-19/Wales/PHWC-491A11/2020\|EPI_ISL_706727\|2020 | ···················· | ···················· | ···················· | ···················· | NNNN················ | ············NNN····· | 1 |
| W | 2021 hCoV-19/Wales/PHWC-491A3F/2020\|EPI_ISL_706729\|2020 | ···················· | ···················· | ···················· | ···················· | NNNNNNNNNNNNNNNNNNNN | NNNNNNNNNNNNNNN····· | 1 |
| W | 2025 hCoV-19/Wales/PHWC-491A7B/2020\|EPI_ISL_706733\|2020 | ···················· | ···················· | ···················· | ···················· | NNNNNNNNNNNNNNNNNNNN | NNNNNNNNNNNNNNNNNNNN | 1 |
| W | 2027 hCoV-19/Wales/PHWC-491A99/2020\|EPI_ISL_706735\|2020 | ···················· | ···················· | ···················· | ···················· | NNNNNNNNNNNNNNNNNNNN | ···················· | 1 |
| W | 2028 hCoV-19/Wales/PHWC-491AA8/2020\|EPI_ISL_706736\|2020 | ···················· | ···················· | ···················· | ···················· | NNNNNNNNNNNNNNNNNNNN | ···················· | 1 |
| W | 2030 hCoV-19/Wales/PHWC-491B96/2020\|EPI_ISL_706751\|2020 | ···················· | ···················· | ···················· | ···················· | NNNNNNNNNNNNNNNNNNNN | NNNNN··············· | 1 |
| W | 2034 hCoV-19/Wales/PHWC-491BD2/2020\|EPI_ISL_706755\|2020 | ···················· | ···················· | ···················· | ···················· | NNNNNNNNNNNN········ | ···················· | 1 |
| W | 2038 hCoV-19/Wales/PHWC-491C2A/2020\|EPI_ISL_706759\|2020 | ···················· | ···················· | ···················· | ···················· | ···················· | NNNNNNNNNNNNNNNNNNNN | 1 |
| W | 2041 hCoV-19/Wales/PHWC-491C75/2020\|EPI_ISL_706762\|2020 | ···················· | ···················· | ···················· | ···················· | NNNNNNNNNNNNNNNNNNNN | NNNNN··············· | 1 |
| W | 2162 hCoV-19/Wales/PHWC-492412/2020\|EPI_ISL_705412\|2020 | ···················· | ···················· | ···················· | ···················· | NNNNNNNNNNNNNNNNNNNN | ···········NNNN·NNNN | 1 |
| W | 2167 hCoV-19/Wales/PHWC-491079/2020\|EPI_ISL_705418\|2020 | ···················· | ···················· | ···················· | ···················· | NNNNNNNNNNNNNNNNNNNN | NNNNNNNNNNNNNNNNNNNN | 1 |
| W | 2169 hCoV-19/Wales/PHWC-491AB7/2020\|EPI_ISL_706737\|2020 | ···················· | ···················· | ···················· | ···················· | NNNNNNNNNNNNNNNNNNNN | NNNNNNNNNNNNNNNNNNNN | 1 |
| W | 2173 hCoV-19/Wales/PHWC-491AF3/2020\|EPI_ISL_706741\|2020 | ···················· | ···················· | ···················· | ···················· | NNNNNNNNNNNNNNNNNNNN | ···················· | 1 |
| W | 2174 hCoV-19/Wales/PHWC-491B0F/2020\|EPI_ISL_706742\|2020 | ···················· | ···················· | ···················· | ···················· | NNNNNNNNNNNNNNNNNNNN | NNNNNNNNNNNNNNNNNNNN | 1 |
| W | 2178 hCoV-19/Wales/PHWC-491B4B/2020\|EPI_ISL_706746\|2020 | ···················· | ·A·················· | ···················· | ···················· | NNNNNNNNNNNNNNNNNNNN | NNNNNNNNNNNNNNNNNNNN | 1 |
| W | 2179 hCoV-19/Wales/PHWC-491B5A/2020\|EPI_ISL_706747\|2020 | ···················· | ···················· | ···················· | ···················· | NNNNNNNNNNNNNNNNNNNN | NNNNNNNNNNNNNNNNNNNN | 1 |
| W | 2180 hCoV-19/Wales/PHWC-491B69/2020\|EPI_ISL_706748\|2020 | ···················· | ···················· | ···················· | ···················· | NNNNNNNNNNNNNNNNNNNN | ····N··············· | 1 |
| W | 2581 hCoV-19/USA/AZ-TG532053/2020\|EPI_ISL_695401\|2020-0 | ···················· | ·············A······ | ···················· | ···················· | ··NNNNNNNNNNNNNNNNNN | ···················· | 1 |
| W | 2585 hCoV-19/USA/AZ-TG532087/2020\|EPI_ISL_695412\|2020-0 | ···················· | ·A·················· | ···················· | ···················· | ··NNNNNNNNNNNNNNNNNN | ···················· | 1 |
| W | 2883 hCoV-19/USA/AZ-TG532045/2020\|EPI_ISL_695398\|2020-0 | ···················· | ·············A······ | ···················· | ···················· | ·····N··NNNNNNNNNNNN | ···················· | 1 |
| W | 3118 hCoV-19/India/MH-ACTREC-371/2020\|EPI_ISL_700020\|20 | ···················· | ···················· | ···················· | ···················· | ·······NNNNNNNNNNNNN | ···················· | 1 |
| W | 3131 hCoV-19/India/MH-ACTREC-384/2020\|EPI_ISL_700033\|20 | ···················· | ···················· | ···················· | ···················· | ········NNNNNNNNNNNN | ···················· | 1 |
| W | 3152 hCoV-19/India/MH-ACTREC-407/2020\|EPI_ISL_700056\|20 | ···················· | ···················· | ···················· | ···················· | ·······NNNNNNNNNNNNN | NNNNNNNNNNNNNNNNNNNN | 1 |
| W | 3169 hCoV-19/India/MH-ACTREC-424/2020\|EPI_ISL_700073\|20 | ···················· | ···················· | ···················· | ···················· | ········NNNNNNNNNNNN | ···················· | 1 |
| W | 3177 hCoV-19/India/MH-ACTREC-433/2020\|EPI_ISL_700082\|20 | ···················· | ···················· | ···················· | ···················· | ···················· | NNNNNNNNNNNNNNNNNNNN | 1 |
| W | 3180 hCoV-19/India/MH-ACTREC-436/2020\|EPI_ISL_700085\|20 | ···················· | ···················· | ···················· | ···················· | ·······NNNNNNNNNNNNN | ···················· | 1 |
| W | 3181 hCoV-19/India/MH-ACTREC-437/2020\|EPI_ISL_700086\|20 | ···················· | ···················· | ···················· | ···················· | ··········NNNNNNNNNN | ···················· | 1 |
| W | 3184 hCoV-19/India/MH-ACTREC-440/2020\|EPI_ISL_700089\|20 | ···················· | ···················· | ···················· | ···················· | ·······NNNNNNNNNNNNN | ···················· | 1 |
| W | 3185 hCoV-19/India/MH-ACTREC-441/2020\|EPI_ISL_700090\|20 | ···················· | ···················· | ···················· | ···················· | ···················· | NNNNNNNNNNNNNNNNNNNN | 1 |
| W | 3186 hCoV-19/India/MH-ACTREC-442/2020\|EPI_ISL_700091\|20 | ···················· | ···················· | ···················· | ···················· | ···················· | NNNNNNNNNNNNN······· | 1 |
| W | 3342 hCoV-19/South Africa/NHLS-UCT-GS-7025-KRISP/2020\|E | ···················· | ···················· | ···················· | ···················· | NNNNNNNNNNNNNNNNNNNN | NNNNNNNNNNNNNNNNNNNN | 1 |
| W | 3343 hCoV-19/South Africa/NHLS-UCT-GS-7035-KRISP/2020\|E | ···················· | ···················· | ···················· | ···················· | ··NNNNNNNNNNNNNNNNNN | NNNNNNNNNNNNNNNNNNNN | 1 |
| W | 3344 hCoV-19/South Africa/NHLS-UCT-GS-7065-KRISP/2020\|E | ···················· | ···················· | ···················· | ···················· | ············NNNNNNNN | NNNNNNNNNNNNNNNNNNNN | 1 |
| W | 3345 hCoV-19/South Africa/NHLS-UCT-GS-7036-KRISP/2020\|E | ···················· | ···················· | ···················· | ···················· | NNNNNNNNNNNNNNNNNNNN | NNNNNNNNNNNNNNNNNNNN | 1 |
| W | 3348 hCoV-19/South Africa/NHLS-UCT-GS-7037-KRISP/2020\|E | ···················· | ···················· | ···················· | ···················· | ·NNNNNNNNNNNNNNNNNNN | NNNNNNNNNNNNNNNNNNNN | 1 |
| W | 3349 hCoV-19/South Africa/NHLS-UCT-GS-7058-KRISP/2020\|E | ···················· | ···················· | ···················· | ···················· | NNNNNNNNNNNNNNNNNNNN | NNNNNNNNNNNNNNNNNNNN | 1 |
| W | 3350 hCoV-19/South Africa/NHLS-UCT-GS-7032-KRISP/2020\|E | ···················· | ···················· | ···················· | ···················· | ·NNNNNNNNNNNNNNNNNNN | NNNNNNNNNNNNNNNNNNNN | 1 |
| W | 3351 hCoV-19/South Africa/NHLS-UCT-GS-7034-KRISP/2020\|E | ···················· | ···················· | ···················· | ···················· | ··NNNNNNNNNNNNNNNNNN | ········NNNNNNNNNNNN | 1 |
| W | 3362 hCoV-19/India/MH-ACTREC-461/2020\|EPI_ISL_700109\|20 | ···················· | ···················· | ···················· | ···················· | ·······NNNNNNNNNNNNN | NNNNNNNNNNNNNNNNNNNN | 1 |
| W | 3363 hCoV-19/India/MH-ACTREC-462/2020\|EPI_ISL_700110\|20 | ···················· | ···················· | ···················· | ···················· | ·······NNNNNNNNNNNNN | NNNNNNNNNNNNNNNNNNNN | 1 |
| W | 3375 hCoV-19/India/MH-ACTREC-474/2020\|EPI_ISL_700122\|20 | ···················· | ···················· | ···················· | ···················· | ········NNNNN······· | ···················· | 1 |
| W | 3380 hCoV-19/India/MH-ACTREC-479/2020\|EPI_ISL_700127\|20 | ···················· | ···················· | ···················· | ···················· | ·······NNNNNNNNNNNNN | ···················· | 1 |
| W | 3384 hCoV-19/India/MH-ACTREC-484/2020\|EPI_ISL_700132\|20 | ···················· | ·A·················· | ···················· | ···················· | ·······NNNNNNNNNNNNN | NNNNNNNNNNNNNNNNNNNN | 1 |
| W | 3430 hCoV-19/India/MH-ACTREC-534/2020\|EPI_ISL_700180\|20 | ···················· | ···················· | ···················· | ···················· | ·······NNNNNNNNNNNNN | ···················· | 1 |
| W | 3449 hCoV-19/India/MH-ACTREC-554/2020\|EPI_ISL_700199\|20 | ···················· | ·R·················· | ···················· | ···················· | ······NNNNNNNNNNNNNN | NNNNNNNNNNNNNNNNNNNN | 1 |
| W | 3713 hCoV-19/Norway/5617/2020\|EPI_ISL_708126\|2020-11-10 | ···················· | ···················· | ···················· | ···················· | NNNNNNNNNNNNNNNNNNNN | NNNNNNNNNNNNNNNNNNNN | 1 |
| W | 3714 hCoV-19/Norway/5618/2020\|EPI_ISL_708127\|2020-11-10 | ···················· | ···················· | ···················· | ···················· | NNNNNNNNNNNNNNNNNNNN | NNNNNNNNNNNNNNNNNNNN | 1 |
| W | 3715 hCoV-19/Norway/5619/2020\|EPI_ISL_708128\|2020-11-10 | ···················· | ···················· | ···················· | ···················· | NNNNNNNNNNN········· | ···················· | 1 |
| W | 3717 hCoV-19/Norway/5621/2020\|EPI_ISL_708130\|2020-11-10 | ···················· | ···················· | ···················· | ···················· | NNNNN·N····NNN······ | ···················· | 1 |
| W | 3975 hCoV-19/England/LIVE-DC6E4B/2020\|EPI_ISL_705927\|20 | ···················· | ···················· | ···················· | ···················· | NNNNNNNNNNNNNNNNNNNN | NNNNNNNNNNNNNNNNNNNN | 1 |
| W | 3976 hCoV-19/England/LIVE-DC7114/2020\|EPI_ISL_705928\|20 | ···················· | ···················· | ···················· | ···················· | NNNNNNNNNNNNNNNNNNNN | NNNNNNNNNNNNNNNNNNNN | 1 |
| W | 3993 hCoV-19/England/LOND-1251AA3/2020\|EPI_ISL_705945\|2 | ···················· | ···················· | ···················· | ···················· | NNNNNNNNNNNNNNNNNNNN | ···················· | 1 |
| W | 3999 hCoV-19/England/LOND-1251B82/2020\|EPI_ISL_705951\|2 | ···················· | ···················· | ···················· | ···················· | NNNNNNNNNNNNNNNNNNNN | NNNNNNNNNNNNNNNNNNNN | 1 |
| W | 4004 hCoV-19/England/LOND-1251E6B/2020\|EPI_ISL_705956\|2 | ···················· | ···················· | ···················· | ···················· | ·NNNNNNNNNNNNNNNNNNN | ···················· | 1 |
| W | 4008 hCoV-19/England/LOND-1252046/2020\|EPI_ISL_705960\|2 | ···················· | ···················· | ···················· | ···················· | ··NNNNNNNNNNNNNNNNNN | NNNNNNNNNNNNNNNNNNNN | 1 |
| W | 4772 hCoV-19/England/NORW-F8BDD/2020\|EPI_ISL_704858\|202 | ···················· | ···················· | ···················· | ···················· | NNNNNNNNNNNNNNNNNNNN | NNNNNNNNNNNNNNNNNNNN | 1 |
| W | 5002 hCoV-19/England/NORW-F61FB/2020\|EPI_ISL_704847\|202 | ···················· | ···················· | ···················· | ···················· | NNNNNNNNNNNNNNNNNNNN | NNNNNNNNNNNNNNNNNNNN | 1 |
| W | 5206 hCoV-19/India/MH-ACTREC-271/2020\|EPI_ISL_699923\|20 | ···················· | ···················· | ···················· | ···················· | ·······NNNNNNNNNNNNN | NNNNNNNNNNNNNNNNNNNN | 1 |
| W | 5320 hCoV-19/Wales/PHWC-490326/2020\|EPI_ISL_704918\|2020 | ···················· | ···················· | ···················· | ···················· | NNNNNNNNNNNNNNNNNNNN | N··················· | 1 |
| W | 5349 hCoV-19/Wales/PHWC-491EBB/2020\|EPI_ISL_706791\|2020 | ···················· | ···················· | ···················· | ···················· | NNNNNNNNNNNNNNNNNNNN | ·················NNN | 1 |
| W | 5350 hCoV-19/Wales/PHWC-491ECA/2020\|EPI_ISL_706792\|2020 | ···················· | ···················· | ···················· | ···················· | NNNNNNNNNNNNNNNNNNNN | ·NNNNNN············· | 1 |
| W | 5351 hCoV-19/Wales/PHWC-491ED9/2020\|EPI_ISL_706793\|2020 | ···················· | ···················· | ···················· | ···················· | NNNNNNN············· | ···················· | 1 |
| W | 5352 hCoV-19/Wales/PHWC-491EE8/2020\|EPI_ISL_706794\|2020 | ···················· | ···················· | ···················· | ···················· | NNNNNNNNNNNNNNNNNNNN | NNNNNNNNNNNNNNNNNNNN | 1 |
| W | 5354 hCoV-19/Wales/PHWC-491F03/2020\|EPI_ISL_706796\|2020 | ···················· | ···················· | ···················· | ···················· | NNNNNNNNNNNNNNNNNNNN | NNNNNNNNNNNNNNNNNNNN | 1 |
| W | 5355 hCoV-19/Wales/PHWC-491F4F/2020\|EPI_ISL_706797\|2020 | ···················· | ···················· | ···················· | ···················· | NNNNNNNNNNNNNNNNNNNN | NNNNNNNNNNNNNNNNNNNN | 1 |
| W | 5458 hCoV-19/India/MH-ACTREC-280/2020\|EPI_ISL_699932\|20 | ···················· | ···················· | ···················· | ···················· | ········NNNNNNNNNNNN | ···················· | 1 |
| W | 5589 hCoV-19/Wales/PHWC-4922BE/2020\|EPI_ISL_705441\|2020 | ···················· | ···················· | ···················· | ···················· | NNNNNNNNNNNNNNNNNNNN | NNNNNNNNNNNNNNN····· | 1 |

| | | | | | | | | | |
|---|---|---|---|---|---|---|---|---|---|
| W | 5616 | hCoV-19/Wales/PHWC-491D54/2020\|EPI_ISL_706774\|2020 | ···················· | ···················· | ···················· | ···················· | NNN················· | NNNNNNNNNNNNNNN····· | 1 |
| W | 5620 | hCoV-19/Wales/PHWC-491DAF/2020\|EPI_ISL_706778\|2020 | ···················· | ···················· | ···················· | ···················· | NNNNNNNNNNNNNNNNNNNN | ···················· | 1 |
| W | 5622 | hCoV-19/Wales/PHWC-491DCD/2020\|EPI_ISL_706780\|2020 | ···················· | ···················· | ···················· | ···················· | ····NNNNNNNNNNNNNNNN | NNNNNN········N·NNNN | 1 |
| W | 5623 | hCoV-19/Wales/PHWC-491DFA/2020\|EPI_ISL_706781\|2020 | ···················· | ···················· | ···················· | ···················· | NNNNNNNNNNNNNNNNNNNN | NNNNNNNNNNNNNNNNNNNN | 1 |
| W | 5625 | hCoV-19/Wales/PHWC-491E15/2020\|EPI_ISL_706783\|2020 | ···················· | ···················· | ···················· | ···················· | NNNNNNNNNNNNNNNNNNNN | NNNNNNNNNNNNNNNNNNNN | 1 |
| W | 5626 | hCoV-19/Wales/PHWC-491E24/2020\|EPI_ISL_706784\|2020 | ···················· | ···················· | ···················· | ···················· | NNNNNNNNNNNNNNNNNNNN | ···················· | 1 |
| W | 5627 | hCoV-19/Wales/PHWC-491E33/2020\|EPI_ISL_706785\|2020 | ···················· | ···················· | ···················· | ···················· | ···················· | NNNNN··············· | 1 |
| W | 5630 | hCoV-19/Wales/PHWC-491E60/2020\|EPI_ISL_706788\|2020 | ···················· | ···················· | ···················· | ···················· | ···················· | ·········NNNNNN····· | 1 |
| W | 5632 | hCoV-19/Wales/PHWC-491F6D/2020\|EPI_ISL_706799\|2020 | ···················· | ···················· | ···················· | ···················· | NNNNNNNNNNNNNNNNNNNN | NNNNNNNNNNNNNNNNNNNN | 1 |
| W | 5671 | hCoV-19/India/MH-ACTREC-253/2020\|EPI_ISL_699905\|20 | ···················· | ···················· | ···················· | ···················· | ·······NNNNNNNNNNNNN | NNNNNNNNNNNNNNNNNNNN | 1 |
| W | 5683 | hCoV-19/India/MH-ACTREC-265/2020\|EPI_ISL_699917\|20 | ···················· | ···················· | ···················· | ···················· | ·······NNNNNNNNNNNNN | NNNNNNNNNNNNNNNNNNNN | 1 |
| W | 5687 | hCoV-19/England/204590575/2020\|EPI_ISL_700652\|2020 | ···················· | ···················· | ···················· | ···················· | NNNNNNNNNNNNNNNNNNNN | ···················· | 1 |
| W | 5777 | hCoV-19/Wales/PHWC-491F8B/2020\|EPI_ISL_706801\|2020 | ···················· | ···················· | ···················· | ···················· | NNNNNNNNNNNNNNNNNNNN | NNNNNNNNNNNNNNNNNNNN | 1 |
| W | 5778 | hCoV-19/Wales/PHWC-491FA9/2020\|EPI_ISL_706802\|2020 | ···················· | ···················· | ···················· | ···················· | NNNNNNNNNNNNNNNNNNNN | NNNNNNNNNNNNNNNNNNNN | 1 |
| W | 5779 | hCoV-19/Wales/PHWC-491FB8/2020\|EPI_ISL_706803\|2020 | ···················· | ···················· | ···················· | ···················· | NNNNNNNNNNNNNNNNNNNN | NNNNNNNNNNNNNNNNNNNN | 1 |
| W | 5786 | hCoV-19/Wales/PHWC-4921C0/2020\|EPI_ISL_706829\|2020 | ···················· | ···················· | ···················· | ···················· | NNNNNNNNNNNNNNNNNNNN | NNNNNNNNNNNNNNNNNNNN | 1 |
| W | 5788 | hCoV-19/Wales/PHWC-492227/2020\|EPI_ISL_706831\|2020 | ···················· | ···················· | ···················· | ···················· | NNNNNNNNNNNNNNNNNNNN | NNNNNNNNNNNNNNNNNNNN | 1 |
| W | 5793 | hCoV-19/Wales/PHWC-4922CD/2020\|EPI_ISL_706836\|2020 | ···················· | ···················· | ···················· | ···················· | NNNNNNNNNNNNNNNNNNNN | NNNNNNNNNNNNNNN····· | 1 |
| W | 5797 | hCoV-19/Wales/PHWC-492360/2020\|EPI_ISL_706840\|2020 | ···················· | ···················· | ···················· | ···················· | NNNNNNNNNNNNNNNNNNNN | NNNNNNNNNNNNNNNNNNNN | 1 |
| W | 5798 | hCoV-19/Wales/PHWC-49238E/2020\|EPI_ISL_706841\|2020 | ···················· | ···················· | ···················· | ···················· | NNNNNNNNNNNNNNNNNNNN | NNNNNNNNNNNNNNNNNNNN | 1 |
| W | 5799 | hCoV-19/Wales/PHWC-4923AC/2020\|EPI_ISL_706842\|2020 | ···················· | ···················· | ···················· | ···················· | NNNNNNNNNNN········· | ···················· | 1 |
| W | 5801 | hCoV-19/Wales/PHWC-4923F7/2020\|EPI_ISL_706844\|2020 | ···················· | ···················· | ···················· | ···················· | NNNNNNNNNNNNNNNNNNNN | NNNNNNNNNNNNNNNNNNNN | 1 |
| W | 5931 | hCoV-19/India/MH-ACTREC-148/2020\|EPI_ISL_699804\|20 | ···················· | ···················· | ···················· | ···················· | ·······NNNNNNNNNNNNN | ···················· | 1 |
| W | 5934 | hCoV-19/India/MH-ACTREC-151/2020\|EPI_ISL_699807\|20 | ···················· | ···················· | ···················· | ···················· | ···················· | NNNNNNNNNNNNNN······ | 1 |
| W | 5937 | hCoV-19/India/MH-ACTREC-155/2020\|EPI_ISL_699811\|20 | ···················· | ···················· | ···················· | ···················· | ·······NNNNNNNNNNNNN | NNNNNNNNNNNNNNNNNNNN | 1 |
| W | 5939 | hCoV-19/India/MH-ACTREC-157/2020\|EPI_ISL_699813\|20 | ···················· | ···················· | ···················· | ···················· | ·······NNNNNNNNNNNNN | NNNNNNNNNNNNNNNNNNNN | 1 |
| W | 5940 | hCoV-19/India/MH-ACTREC-159/2020\|EPI_ISL_699815\|20 | ···················· | ···················· | ···················· | ···················· | ·······NNNNNNNNNNNNN | ···················· | 1 |
| W | 5941 | hCoV-19/India/MH-ACTREC-160/2020\|EPI_ISL_699816\|20 | ···················· | ···················· | ···················· | ···················· | ·······NNNNNNNNNNNNN | NNNNNNNNNNNNNNNNNNNN | 1 |
| W | 5946 | hCoV-19/India/MH-ACTREC-168/2020\|EPI_ISL_699824\|20 | ···················· | ···················· | ···················· | ···················· | ·······NNNNNNNNNNNNN | NNNNNNNNNNNNNNNNNNNN | 1 |
| W | 5948 | hCoV-19/India/MH-ACTREC-173/2020\|EPI_ISL_699828\|20 | ···················· | ···················· | ···················· | ···················· | ·······NNNNNNNNNNNN· | ···················· | 1 |
| W | 5949 | hCoV-19/India/MH-ACTREC-174/2020\|EPI_ISL_699829\|20 | ···················· | ···················· | ···················· | ···················· | ·······NNNNNNNNNNNNN | ···················· | 1 |
| W | 5951 | hCoV-19/India/MH-ACTREC-177/2020\|EPI_ISL_699832\|20 | ···················· | ················T··· | ···················· | ···················· | ·······NNNNNNNNNNNNN | NNNNNNNNNNNNNNNNNNNN | 1 |
| W | 5953 | hCoV-19/India/MH-ACTREC-179/2020\|EPI_ISL_699834\|20 | ···················· | ················T··· | ···················· | ···················· | ···················· | ····NNNNNNNNNNNNNNNN | 1 |
| W | 5959 | hCoV-19/India/MH-ACTREC-189/2020\|EPI_ISL_699843\|20 | ···················· | ···················· | ···················· | ···················· | ·······NNNNNNNNNNNNN | NNNNNNNNNNNNNNNNNNNN | 1 |
| W | 6038 | hCoV-19/England/CAMC-BBBD20/2020\|EPI_ISL_705567\|20 | ···················· | ···················· | ···················· | ···················· | NNNNNNNNNNNNNNNN···· | NNNNNNNNNNNNNNN····· | 1 |
| W | 6047 | hCoV-19/England/CAMC-BBBF39/2020\|EPI_ISL_705576\|20 | ···················· | ···················· | ···················· | ···················· | NNNNNNNNNNNNN······· | ·················NNN | 1 |
| W | 6071 | hCoV-19/Wales/PHWC-49204B/2020\|EPI_ISL_706810\|2020 | ···················· | ·········T·········· | ···················· | ···················· | ···················· | NNNNNNNNNNNNNNN····· | 1 |
| W | 6075 | hCoV-19/Wales/PHWC-492096/2020\|EPI_ISL_706814\|2020 | ···················· | ···················· | ···················· | ···················· | NNNNNNNNNNNNNNNNNNNN | NNNNNNNNNNNNNNNNNNNN | 1 |
| W | 6076 | hCoV-19/Wales/PHWC-4920A5/2020\|EPI_ISL_706815\|2020 | ···················· | ···················· | ···················· | ···················· | NNNNNNNNNNNNNNNNNNNN | N··················· | 1 |
| W | 6081 | hCoV-19/Wales/PHWC-49210C/2020\|EPI_ISL_706820\|2020 | ···················· | ···················· | ···················· | ···················· | NNNNNNNNNNNNNNNNNNNN | NNNNNNNNNNNNNNNNNNNN | 1 |
| W | 6084 | hCoV-19/Wales/PHWC-492139/2020\|EPI_ISL_706823\|2020 | ···················· | ···················· | ···················· | ···················· | NNNNNNNNNNN········· | NNNNN··············· | 1 |
| W | 6090 | hCoV-19/Wales/PHWC-49249A/2020\|EPI_ISL_706850\|2020 | ···················· | ···················· | ···················· | ···················· | NNNNNNNNNNNNNNNNNNNN | ···················· | 1 |
| W | 6093 | hCoV-19/Wales/PHWC-492500/2020\|EPI_ISL_706853\|2020 | ···················· | ···················· | ···················· | ···················· | NNNNNNNNNNNNNNNNNNNN | NNNNNNNNNNNNNNNNNNNN | 1 |
| W | 6099 | hCoV-19/Wales/PHWC-492588/2020\|EPI_ISL_706859\|2020 | ···················· | ···················· | ···················· | ···················· | ···················· | NNNNN·······NNN····· | 1 |
| W | 6103 | hCoV-19/Wales/PHWC-4925C4/2020\|EPI_ISL_706863\|2020 | ···················· | ···················· | ···················· | ···················· | NNNNNNNNNNNNNNNNNNNN | NNNNNNNNNNNNNNN····· | 1 |
| W | 6238 | hCoV-19/India/MH-ACTREC-190/2020\|EPI_ISL_699844\|20 | ···················· | ···················· | ···················· | ···················· | ·······NNNNNNNNNNNNN | NNNNNNNNNNNNNNNNNNNN | 1 |
| W | 6239 | hCoV-19/India/MH-ACTREC-191/2020\|EPI_ISL_699845\|20 | ···················· | ···················· | ···················· | ···················· | ·······NNNNNNNNNNNNN | NNNNNNNNNNNNNNNNNNNN | 1 |
| W | 6241 | hCoV-19/India/MH-ACTREC-196/2020\|EPI_ISL_699850\|20 | ···················· | ·A·················· | ···················· | ···················· | ·······NNNNNNNNNNNNN | ···A················ | 1 |
| W | 6242 | hCoV-19/India/MH-ACTREC-197/2020\|EPI_ISL_699851\|20 | ···················· | ···················· | ···················· | ···················· | ·······NNNNNNNNNNNNN | NNNNNNNNNNNNNNNNNNNN | 1 |
| W | 6246 | hCoV-19/India/MH-ACTREC-201/2020\|EPI_ISL_699855\|20 | ···················· | ···················· | ···················· | ···················· | ·······NNNNNNNNNNNNN | ···················· | 1 |
| W | 6254 | hCoV-19/India/MH-ACTREC-209/2020\|EPI_ISL_699863\|20 | ···················· | ···················· | ···················· | ···················· | ·······NNNNNNNNNNNNN | NNNNNNNNNNNNNNNNNNNN | 1 |
| W | 6326 | hCoV-19/England/QEUH-BF19C0/2020\|EPI_ISL_709107\|20 | ···················· | NNNNNNNNNNNNNNNNNNNN | ···················· | ···················· | ···················· | ···················· | 1 |
| W | 6392 | hCoV-19/USA/CO-CDPHE-2004220691/2020\|EPI_ISL_71018 | ···················· | ···················· | ···················· | ···················· | NNNNNNNNNNNNNNNNNNNN | NNNNNNNNNNNNNNNNNNNN | 1 |
| W | 6393 | hCoV-19/USA/CO-CDPHE-2004220697/2020\|EPI_ISL_71018 | ···················· | ···················· | ···················· | ···················· | NNNNNNNNNNNNNNNNNNNN | NNNNNNNNNNNNNNNNNNNN | 1 |
| W | 6395 | hCoV-19/USA/CO-CDPHE-2004220732/2020\|EPI_ISL_71018 | ···················· | ···················· | ···················· | ···················· | NNNNNNNNNNNNNNNNNNNN | NNNNNNNNNNNNNNNNNNNN | 1 |
| W | 6460 | hCoV-19/India/MH-ACTREC-115/2020\|EPI_ISL_699771\|20 | ···················· | ···················· | ···················· | ···················· | ·······NNNNNNNNNNNNN | NNNNNNNNNNNNNNNNNNNN | 1 |
| W | 6466 | hCoV-19/India/MH-ACTREC-123/2020\|EPI_ISL_699779\|20 | ···················· | ···················· | ···················· | ···················· | ········NNNNNNNNNNNN | NNNNNNNNNNNNNNNNNNNN | 1 |
| W | 6469 | hCoV-19/India/MH-ACTREC-126/2020\|EPI_ISL_699782\|20 | ···················· | ···················· | ···················· | ···················· | ·······NNNNNNNNNNNNN | ··············NNNNNN | 1 |
| W | 6485 | hCoV-19/India/MH-ACTREC-142/2020\|EPI_ISL_699798\|20 | ···················· | NNNNNNNNNNNNNNNNNNNN | ···················· | ···················· | ···················· | ···················· | 1 |
| W | 6649 | hCoV-19/USA/CO-CDPHE-2004220763/2020\|EPI_ISL_71019 | ···················· | ···················· | ···················· | ···················· | NNNNNNNNNNNNNNNNNNNN | NNNNNNNNNNNNNNNNNNNN | 1 |
| W | 6653 | hCoV-19/USA/CO-CDPHE-2004220779/2020\|EPI_ISL_71019 | ···················· | ···················· | ···················· | ···················· | NNNNNNNNNNNNNNNNNNNN | NNNNNNNNNNNNNNNNNNNN | 1 |
| W | 6654 | hCoV-19/USA/CO-CDPHE-2004220780/2020\|EPI_ISL_71019 | ···················· | ···················· | ···················· | ···················· | NNNNNNNNNNNNNNNNNNNN | NNNNNNNNNNNNNNNNNNNN | 1 |
| W | 6717 | hCoV-19/India/MH-ACTREC-049/2020\|EPI_ISL_699705\|20 | ···················· | ···················· | ···················· | ···················· | ···················· | NNNNNNNNNNNNNNNNNNNN | 1 |
| W | 6718 | hCoV-19/India/MH-ACTREC-050/2020\|EPI_ISL_699706\|20 | ···················· | ···················· | ···················· | ···················· | ·······NNNNNNNNNNNNN | ···················· | 1 |
| W | 6721 | hCoV-19/India/MH-ACTREC-055/2020\|EPI_ISL_699711\|20 | ···················· | ···················· | ···················· | ···················· | ·······NNNNNNNNNNNNN | NNNNNNNNNNNNNNNNNNNN | 1 |
| W | 6732 | hCoV-19/India/MH-ACTREC-067/2020\|EPI_ISL_699723\|20 | ···················· | ···················· | ···················· | ···················· | ·······NNNNNNNNNNNNN | NNNNNNNNNNNNNNNNNNNN | 1 |
| W | 6742 | hCoV-19/India/MH-ACTREC-077/2020\|EPI_ISL_699733\|20 | ···················· | ···················· | ···················· | ···················· | ·······NNNNNNNNNNNNN | ···················· | 1 |
| W | 6749 | hCoV-19/India/MH-ACTREC-084/2020\|EPI_ISL_699740\|20 | ···················· | ···················· | ···················· | ···················· | ········NNNNNNNNNNNN | ···················· | 1 |
| W | 6768 | hCoV-19/India/MH-ACTREC-105/2020\|EPI_ISL_699761\|20 | ···················· | ···················· | ···················· | ···················· | ·······NNNNNNNNNNNNN | NNNNNNNNNNNNNNNNNNNN | 1 |
| W | 6890 | hCoV-19/USA/CO-CDPHE-2004230195/2020\|EPI_ISL_71020 | ···················· | ···················· | ···················· | ···················· | NNNNNNNNNNNNNNNNNNNN | NNNNNNNNNNNNNNNNNNNN | 1 |
| W | 6893 | hCoV-19/USA/CO-CDPHE-2004230233/2020\|EPI_ISL_71020 | ···················· | ···················· | ···················· | ···················· | NNNNNNNNNNNNNNNNNNNN | NNNNNNNNNNNNNNNNNNNN | 1 |
| W | 6894 | hCoV-19/USA/CO-CDPHE-2004300317/2020\|EPI_ISL_71020 | ···················· | ···················· | ···················· | ···················· | NNNNNNNNNNNNNNNNNNNN | NNNNNNNNNNNNNNNNNNNN | 1 |
| W | 6895 | hCoV-19/USA/CO-CDPHE-2005060168/2020\|EPI_ISL_71021 | ···················· | ···················· | ···················· | ···················· | NNNNNNNNNNNNNNNNNNNN | NNNNNNNNNNNNNNNNNNNN | 1 |
| W | 7009 | hCoV-19/India/MH-ACTREC-025/2020\|EPI_ISL_699681\|20 | ···················· | ···················· | ···················· | ···················· | ·······NNNN········· | ···················· | 1 |
| W | 7017 | hCoV-19/India/MH-ACTREC-033/2020\|EPI_ISL_699689\|20 | ···················· | ···················· | ···················· | ···················· | ···················· | NNNNNNNNNNNNNNNNNNNN | 1 |
| W | 7084 | hCoV-19/Scotland/EDB10221/2020\|EPI_ISL_705734\|2020 | ···················· | ···················· | ···················· | ···················· | NNNNNNNNNNNNNNNNNNNN | NNNNNNNNNNNNNNNNNNNN | 1 |
| W | 7091 | hCoV-19/Scotland/EDB1031/2020\|EPI_ISL_705741\|2020- | ···················· | ···················· | ···················· | ···················· | NNNNNNNNNNNNNNNNNNNN | NNNNNNNNNNNNNNNNNNNN | 1 |
| W | 7092 | hCoV-19/Scotland/EDB10313/2020\|EPI_ISL_705742\|2020 | ···················· | ···················· | ···················· | ···················· | NNNNNNNNNNNNNNNNNNNN | NNNNNNNNNNNNNNNNNNNN | 1 |
| W | 7107 | hCoV-19/Scotland/EDB10435/2020\|EPI_ISL_705757\|2020 | ···················· | ···················· | ···················· | ···················· | NNNNNNNNNNNNNNNNNNNN | NNNNNNNNNNNNNNNNNNNN | 1 |
| W | 7138 | hCoV-19/England/QEUH-BF1975/2020\|EPI_ISL_709288\|20 | ···················· | NNNNNNNNNNNNNNNNNNNN | ···················· | ···················· | ···················· | ···················· | 1 |

| | | | | | | | | | |
|---|---|---|---|---|---|---|---|---|---|
| W | 7148 | hCoV-19/USA/CO-CDPHE-2004220878/2020\|EPI_ISL_71020 | ···················· | ···················· | ···················· | ···················· | NNNNNNNNNNNNNNNNNNNN | ···················· | 1 |
| W | 7149 | hCoV-19/USA/CO-CDPHE-2004230010/2020\|EPI_ISL_71020 | ···················· | ···················· | ···················· | ···················· | NNNNNNNNNNNNNNNNNNNN | ·NNNNNNNNNNNNNN····· | 1 |
| W | 7150 | hCoV-19/USA/CO-CDPHE-2004230143/2020\|EPI_ISL_71020 | ···················· | ···················· | ···················· | ···················· | NNNNNNNNNNNNNNNNNNNN | NNNNNNNNNNNNNNNNNNNN | 1 |
| W | 7165 | hCoV-19/USA/CO-CDPHE-2011020168/2020\|EPI_ISL_71024 | ···················· | ···················· | ···················· | ···················· | NNNNNNNNNNNNNNNNNNNN | ···················· | 1 |
| W | 7188 | hCoV-19/USA/AZ-TG396024/2020\|EPI_ISL_694798\|2020-0 | ···················· | ···················· | ···················· | ···················· | NNNNNNNNNNNNNNNNNNNN | ···················· | 1 |
| W | 7322 | hCoV-19/Scotland/EDB10516/2020\|EPI_ISL_705774\|2020 | ···················· | ···················· | ···················· | ···················· | NNNNNNNNNNNNNNNNNNNN | NNNNNNNNNNNNNNNNNNNN | 1 |
| W | 7324 | hCoV-19/Scotland/EDB10518/2020\|EPI_ISL_705776\|2020 | ···················· | ···················· | ···················· | ···················· | NNNNNNNNNNNNNNNNNNNN | NNNNNNNNNNNNNNNNNNNN | 1 |
| W | 7335 | hCoV-19/Scotland/EDB740/2020\|EPI_ISL_705796\|2020-0 | ···················· | ·A·················· | ···················· | ···················· | NNNNNNNNNNNNNNNNNNNN | NNNNNNNNNNNNNNNNNNNN | 1 |
| W | 7341 | hCoV-19/Germany/SL-SU-20239546/2020\|EPI_ISL_708013 | ···················· | ···················· | ···················· | ···················· | ···················· | NNNNNNNNNNNNNNNNNN·· | 1 |
| W | 7395 | hCoV-19/USA/CO-CDPHE-2005270144/2020\|EPI_ISL_71031 | ···················· | ···················· | ···················· | ···················· | ·NNNNNNNNNNNNNNNNNNN | ···················· | 1 |
| W | 7406 | hCoV-19/USA/CO-CDPHE-2009040498/2020\|EPI_ISL_71033 | ···················· | ···················· | ···················· | ···················· | ·NNNNNNNNNNN········ | ···················· | 1 |
| W | 7407 | hCoV-19/USA/CO-CDPHE-2009042404/2020\|EPI_ISL_71033 | ···················· | ·A·················· | ···················· | ···················· | ··N················· | NNNNNNNN············ | 1 |
| W | 7414 | hCoV-19/USA/CO-CDPHE-2009170073/2020\|EPI_ISL_71033 | ···················· | ·A·················· | ···················· | ···················· | ·NNNNNNNNNNNNNNNNNNN | NNNNNNNNNNNNNNNNNNNN | 1 |
| W | 7423 | hCoV-19/USA/CO-CDPHE-2009184258/2020\|EPI_ISL_71034 | ···················· | ···················· | ···················· | ···················· | NNNNNNNNNNNNNNNNNNNN | NNNNNNNNNNNNNNNNNNNN | 1 |
| W | 7488 | hCoV-19/Scotland/EDB9335/2020\|EPI_ISL_705800\|2020- | ···················· | ···················· | ···················· | ···················· | NNNNNNNNNNNNNNNNNNNN | NNNNNNNNNNNNNNNNNNNN | 1 |
| W | 7491 | hCoV-19/Scotland/EDB9785/2020\|EPI_ISL_705803\|2020- | ···················· | ···················· | ···················· | ···················· | NNNNNNNNNNNNNNNNNNNN | NNNNNNNNNNNNNNNNNNNN | 1 |
| W | 7523 | hCoV-19/England/LIVE-DC2AE1/2020\|EPI_ISL_705836\|20 | ···················· | ···················· | ···················· | ···················· | ·······NNNNNNNNNNNNN | ···················· | 1 |
| W | 7531 | hCoV-19/England/LIVE-DC3998/2020\|EPI_ISL_705865\|20 | ···················· | ···················· | ···················· | ···················· | NNNNNNNNNNNNNNNNNNNN | NNNNNNNNNNNNNNNNNNNN | 1 |
| W | 7533 | hCoV-19/England/LIVE-DC3A4A/2020\|EPI_ISL_705867\|20 | ···················· | ···················· | ···················· | ···················· | NNNNNNNNNNNNNNNNNNNN | NNNNNNNNNNNNNNNNNNNN | 1 |
| W | 7534 | hCoV-19/England/LIVE-DC3A59/2020\|EPI_ISL_705868\|20 | ···················· | ········T··········· | ···················· | ···················· | NNNNNNNNNNNNNNNNNNNN | NNNNNNNNNNNNNNNNNNNN | 1 |
| W | 7637 | hCoV-19/USA/CO-CDPHE-2008063260/2020\|EPI_ISL_71032 | ···················· | ···················· | ···················· | ···················· | NNNNNNNNNNNNNNNNNNNN | ···················· | 1 |
| W | 7641 | hCoV-19/USA/CO-CDPHE-2008201602/2020\|EPI_ISL_71032 | ···················· | ···················· | ···················· | ···················· | NNNNNNNNNNNNNNNNNNNN | NNNNNNNNNNNNNNN····· | 1 |
| W | 7646 | hCoV-19/USA/CO-CDPHE-2010135868/2020\|EPI_ISL_71035 | ···················· | ···················· | ···················· | ···················· | NNNNNNNNNNNNNNNNNNNN | NNNNNNNNNNNNNNN····· | 1 |
| W | 7655 | hCoV-19/USA/CO-CDPHE-2010280196/2020\|EPI_ISL_71036 | ···················· | ···················· | ···················· | ···················· | ··NNNNNNNNNNNNNNNNNN | NNNNNNNNNNNNNNN····· | 1 |
| W | 7661 | hCoV-19/USA/CO-CDPHE-2010280224/2020\|EPI_ISL_71036 | ···················· | ···················· | ···················· | ···················· | NNNNNNNNNNNNNNNNNNNN | NNNNNNNNNNNNNNNNNNNN | 1 |
| W | 7821 | hCoV-19/England/LIVE-DC3BFC/2020\|EPI_ISL_705872\|20 | ···················· | ···················· | ···················· | ···················· | NNNNNNNNNNNNNNNNNNNN | NNNNNNNNNNNNNNNNNNNN | 1 |
| W | 7822 | hCoV-19/England/LIVE-DC3C62/2020\|EPI_ISL_705873\|20 | ···················· | ···················· | ···················· | ···················· | NNNNNNNNNNNNNNNNNNNN | NNNNNNNNNNNNNNNNNNNN | 1 |
| W | 7823 | hCoV-19/England/LIVE-DC3C80/2020\|EPI_ISL_705874\|20 | ···················· | ···················· | ···················· | ···················· | NNNNNNNNNNNNNNNNNNNN | NNNNNNNNNNNNNNNNNNNN | 1 |
| W | 7824 | hCoV-19/England/LIVE-DC3CAE/2020\|EPI_ISL_705875\|20 | ···················· | ········T··········· | ···················· | ···················· | NNNNNNNNNNNNNNNNNNNN | NNNNNNNNNNNNNNNNNNNN | 1 |
| W | 7826 | hCoV-19/England/LIVE-DC3D14/2020\|EPI_ISL_705877\|20 | ···················· | ···················· | ···················· | ···················· | NNNNNNNNNNNNNNNNNNNN | NNNNNNNNNNNNNNNNNNNN | 1 |
| X | 78 | hCoV-19/England/QEUH-BA067E/2020\|EPI_ISL_719642\|20 | ···················· | NNNNNNNNNNNNNNNNNNNN | ···················· | ···················· | ···················· | ···A················ | 1 |
| X | 103 | hCoV-19/England/QEUH-BA09DF/2020\|EPI_ISL_719677\|20 | ···················· | NNNNNNNNNNNNNNNNNNNN | ···················· | ···················· | ···················· | ···················· | 1 |
| X | 495 | hCoV-19/Scotland/EDB10822/2020\|EPI_ISL_724240\|2020 | ···················· | ···················· | ···················· | ···················· | NNNNNNNNNNNNNNNNNNNN | NNNNNNNNNNNNNNNNNNNN | 1 |
| X | 648 | hCoV-19/England/QEUH-B9ECAE/2020\|EPI_ISL_719727\|20 | ···················· | NNNNNNNNNNNNNNNNNNNN | ···················· | ···················· | ···················· | ···················· | 1 |
| X | 655 | hCoV-19/England/QEUH-BA05F9/2020\|EPI_ISL_719734\|20 | ···················· | NNNNNNNNNNNNNNNNNNNN | ···················· | ···················· | ···················· | ···················· | 1 |
| X | 1449 | hCoV-19/USA/MN-MDH-2169/2020\|EPI_ISL_723120\|2020-1 | ···················· | ···················· | ···················· | ···················· | ·····NNNNNN········· | ···················· | 1 |
| X | 2606 | hCoV-19/Belgium/UGent-267/2020\|EPI_ISL_718005\|2020 | ···················· | ···················· | ···················· | ···················· | NNNNNNNNNNNNNNNNNNNN | NNNNNNNNNNNNNNNNNNNN | 1 |
| X | 2609 | hCoV-19/Belgium/UGent-271/2020\|EPI_ISL_718008\|2020 | ···················· | ···················· | ···················· | ···················· | NNNNNNNNNNNNNNNNNNNN | NNNNNNNNNNNNNNNNNNNN | 1 |
| X | 2611 | hCoV-19/Belgium/UGent-274/2020\|EPI_ISL_718010\|2020 | ···················· | ···················· | ···················· | ···················· | NNNNNNNNNNNNNNNNNNNN | NNNNNNNNNNNNNNNNNNNN | 1 |
| X | 2614 | hCoV-19/Belgium/UGent-278/2020\|EPI_ISL_718013\|2020 | ···················· | ···················· | ···················· | ···················· | NNNNNNNNNNNNNNNNNNNN | NNNNNNNNNNNNNNNNNNNN | 1 |
| X | 2620 | hCoV-19/Belgium/UGent-290/2020\|EPI_ISL_718019\|2020 | ···················· | ···················· | ···················· | ···················· | NNNNNNNNNNNNNNNNNNNN | NNNNNNNNNNNNNNNNNNNN | 1 |
| X | 2623 | hCoV-19/Belgium/UGent-293/2020\|EPI_ISL_718022\|2020 | ···················· | ···················· | ···················· | ···················· | NNNNNNNNNNNNNNNNNNNN | NNNNNNNNNNNNNNNNNNNN | 1 |
| X | 2742 | hCoV-19/Italy/LAZ-INMI-101/2020\|EPI_ISL_721624\|202 | ···················· | ···················· | ···················· | ···················· | NNNNNNNNNNNNNNN····· | ···················· | 1 |
| X | 2746 | hCoV-19/Italy/LAZ-INMI-105/2020\|EPI_ISL_721628\|202 | ···················· | ···················· | ···················· | ···················· | NNNNNNNNNNNNNNN····· | ···················· | 1 |
| X | 4482 | hCoV-19/Brazil/RJ-00539/2020\|EPI_ISL_717923\|2020-1 | ···················· | ···················· | ···················· | ···················· | ·······NNNNNNNNNNNNN | NNNNNNNNNNNNNNNNNNNN | 1 |
| X | 4484 | hCoV-19/Brazil/RJ-00541/2020\|EPI_ISL_717925\|2020-1 | ···················· | ···················· | ···················· | ···················· | NNNNNNNNNNNNNNNNNNNN | NNNNNNNNNNNNNNNNNNNN | 1 |
| X | 4539 | hCoV-19/Belgium/UGent-255/2020\|EPI_ISL_717995\|2020 | ···················· | ···················· | ···················· | ···················· | NNNNNNNNNNNNNNNNNNNN | NNNNNNNNNNNNNNNNNNNN | 1 |
| X | 4630 | hCoV-19/Italy/LAZ-INMI-94/2020\|EPI_ISL_710544\|2020 | ···················· | ···················· | ···················· | ···················· | NNNNNNNNNNNNNNN····· | ···················· | 1 |
| X | 5138 | hCoV-19/Scotland/QEUH-BF1A18/2020\|EPI_ISL_710650\|2 | ···················· | NNNNNNNNNNNNNNNNNNNN | ···················· | ···················· | ···················· | ···················· | 1 |
| X | 6770 | hCoV-19/Belgium/UGent-198/2020\|EPI_ISL_717604\|2020 | ···················· | ···················· | ···················· | ···················· | NNNNNNNNNNNNNNNNNNNN | NNNNNNNNNNNNNNNNNNNN | 1 |
| X | 6771 | hCoV-19/Belgium/UGent-218/2020\|EPI_ISL_717605\|2020 | ···················· | ···················· | ···················· | ···················· | NNNNNNNNNNNNNNNNNNNN | NNNNNNNNNNNNNNNNNNNN | 1 |
| X | 6772 | hCoV-19/Belgium/UGent-149/2020\|EPI_ISL_717606\|2020 | ···················· | ··················C· | ···················· | ···················· | NNNNNNNNNNNNNNNNNNNN | NNNNNNNNNNNNNNNNNNNN | 1 |
| X | 6773 | hCoV-19/Belgium/UGent-191/2020\|EPI_ISL_717607\|2020 | ···················· | ···················· | ···················· | ···················· | NNNNNNNNNNNNNNNNNNNN | NNNNNNNNNNNNNNNNNNNN | 1 |
| X | 6774 | hCoV-19/Belgium/UGent-192/2020\|EPI_ISL_717608\|2020 | ···················· | ···················· | ···················· | ···················· | NNNNNNNNNNNNNNNNNNNN | NNNNNNNNNNNNNNNNNNNN | 1 |
| X | 6775 | hCoV-19/Belgium/UGent-194/2020\|EPI_ISL_717609\|2020 | ···················· | ···················· | ···················· | ···················· | NNNNNNNNNNNNNNNNNNNN | NNNNNNNNNNNNNNNNNNNN | 1 |
| X | 6776 | hCoV-19/Belgium/UGent-202/2020\|EPI_ISL_717610\|2020 | ···················· | ···················· | ···················· | ···················· | NNNNNNNNNNNNNNNNNNNN | NNNNNNNNNNNNNNNNNNNN | 1 |
| X | 6777 | hCoV-19/Belgium/UGent-203/2020\|EPI_ISL_717611\|2020 | ···················· | ···················· | ···················· | ···················· | NNNNNNNNNNNNNNNNNNNN | NNNNNNNNNNNNNNNNNNNN | 1 |
| X | 6781 | hCoV-19/Belgium/UGent-215/2020\|EPI_ISL_717615\|2020 | ···················· | ···················· | ···················· | ···················· | NNNNNNNNNNNNNNNNNNNN | NNNNNNNNNNNNNNNNNNNN | 1 |
| X | 6782 | hCoV-19/Belgium/UGent-216/2020\|EPI_ISL_717616\|2020 | ···················· | ···················· | ···················· | ···················· | NNNNNNNNNNNNNNNNNNNN | NNNNNNNNNNNNNNNNNNNN | 1 |
| X | 6783 | hCoV-19/Belgium/UGent-217/2020\|EPI_ISL_717617\|2020 | ···················· | ···················· | ···················· | ···················· | NNNNNNNNNNNNNNNNNNNN | NNNNNNNNNNNNNNNNNNNN | 1 |
| X | 6784 | hCoV-19/Belgium/UGent-219/2020\|EPI_ISL_717618\|2020 | ···················· | ···················· | ···················· | ···················· | NNNNNNNNNNNNNNNNNNNN | NNNNNNNNNNNNNNNNNNNN | 1 |
| X | 6785 | hCoV-19/Belgium/UGent-220/2020\|EPI_ISL_717619\|2020 | ···················· | ···················· | ···················· | ···················· | NNNNNNNNNNNNNNNNNNNN | NNNNNNNNNNNNNNNNNNNN | 1 |
| X | 7103 | hCoV-19/Spain/AR-IBV-98010657/2020\|EPI_ISL_722267\| | ···················· | ···················· | ···················· | ···················· | NNNNNNNNNNNNNNNNNNNN | ···················· | 1 |
| X | 7108 | hCoV-19/Spain/AR-IBV-98010737/2020\|EPI_ISL_722272\| | ···················· | ···················· | ···················· | ···················· | NNNNNNNNNNNN········ | ···················· | 1 |
| Y | 93 | hCoV-19/Finland/4CF7/2020\|EPI_ISL_732613\|2020-12-0 | ···················· | ·A·················· | ···················· | ···················· | ·····NNNN··········· | ···················· | 1 |
| Y | 308 | hCoV-19/Wales/PHWC-497310/2020\|EPI_ISL_726812\|2020 | ···················· | ···················· | ···················· | ···················· | NNNNNNNNNNNNNNNNNNNN | NNNNNNNNNNNNNNNNNNNN | 1 |
| Y | 309 | hCoV-19/Wales/PHWC-49732F/2020\|EPI_ISL_726813\|2020 | ···················· | ···················· | ···················· | ···················· | NNNNNNNNNNNNNNNNN··· | ···················· | 1 |
| Y | 310 | hCoV-19/Wales/PHWC-49733E/2020\|EPI_ISL_726814\|2020 | ···················· | ···················· | ···················· | ···················· | NNNNNNNNNNNNNNNNNNNN | NNNNNNNNNNNNNNN·NNNN | 1 |
| Y | 323 | hCoV-19/Wales/PHWC-49740E/2020\|EPI_ISL_726827\|2020 | ···················· | ···················· | ···················· | ···················· | NNNNNNNNNNNNNNNNNNNN | NNNNNNNNNNNNNNN····· | 1 |
| Y | 325 | hCoV-19/Wales/PHWC-49742C/2020\|EPI_ISL_726829\|2020 | ···················· | ···················· | ···················· | ···················· | NNNNNNNNNNNNNNNNN··· | NNNNNNNNNNN········· | 1 |
| Y | 342 | hCoV-19/Wales/PHWC-497565/2020\|EPI_ISL_726847\|2020 | ···················· | ···················· | ···················· | ···················· | NNNNNNNNNNNNNNNNNNNN | ··NNNNNNNNNNNNN····· | 1 |
| Y | 344 | hCoV-19/Wales/PHWC-497583/2020\|EPI_ISL_726849\|2020 | ···················· | ···················· | ···················· | ···················· | NNNNNNNNNNNNNNNNNNNN | ···················· | 1 |
| Y | 348 | hCoV-19/Wales/PHWC-4975ED/2020\|EPI_ISL_726854\|2020 | ···················· | ···················· | ···················· | ···················· | NNNNNNNNNNNNNNNNNNNN | ···················· | 1 |
| Y | 352 | hCoV-19/Wales/PHWC-497644/2020\|EPI_ISL_726860\|2020 | ···················· | ···················· | ···················· | ···················· | NNNNNNNNNNNNNNNNNNNN | ···················· | 1 |
| Y | 363 | hCoV-19/Wales/PHWC-49776F/2020\|EPI_ISL_726876\|2020 | ···················· | ···················· | ···················· | ···················· | ···················· | NNNNNNNNNNNNNNN····· | 1 |
| Y | 368 | hCoV-19/Wales/PHWC-4977BA/2020\|EPI_ISL_726881\|2020 | ···················· | ···················· | ···················· | ···················· | ·········NNNNNNNNNNN | ···················· | 1 |
| Y | 514 | hCoV-19/Wales/PHWC-49793C/2020\|EPI_ISL_726905\|2020 | ···················· | ···················· | ···················· | ···················· | NNNNNNNNNNNNNNNNNNNN | NNNNNNNNNNNNNNNNNNNN | 1 |
| Y | 523 | hCoV-19/Wales/PHWC-4979C3/2020\|EPI_ISL_726914\|2020 | ···················· | ···················· | ···················· | ···················· | ···················· | NNNNNNNNNNNNNNN····· | 1 |
| Y | 531 | hCoV-19/Switzerland/AG-ETHZ-400041/2020\|EPI_ISL_72 | ···················· | ···················· | ···················· | ···················· | ·······NNNNNNNNNNNNN | ···················· | 1 |
| Y | 727 | hCoV-19/Spain/RI-IBV-99010903/2020\|EPI_ISL_732749\| | ···················· | ···················· | ···················· | ···················· | ·NNNNNN····NNNNNNNNN | ···················· | 1 |

| | | | | | | | | | |
|---|---|---|---|---|---|---|---|---|---|
| Y | 728 | hCoV-19/Spain/RI-IBV-99010904/2020\|EPI_ISL_732750\| | ···················· | ···················· | ···················· | ···················· | NNNNNNNNNNNNNNNNNNNN | ···················· | 1 |
| Y | 729 | hCoV-19/Spain/RI-IBV-99010905/2020\|EPI_ISL_732751\| | ···················· | ···················· | ···················· | ···················· | ·NNNNNN············· | NNNNNNNNNNNNNNNNNNNN | 1 |
| Y | 730 | hCoV-19/Spain/RI-IBV-99010906/2020\|EPI_ISL_732752\| | ···················· | ···················· | ···················· | ···················· | NNNNNNNNNNNNNNNNNNNN | NNNNNNNNNNNNNNNNNNNN | 1 |
| Y | 731 | hCoV-19/Spain/RI-IBV-99010907/2020\|EPI_ISL_732753\| | ···················· | ···················· | ···················· | ···················· | NNNNNNNNNNNNNNNNNNNN | NNNNNNNNNNNNNNNNNNNN | 1 |
| Y | 732 | hCoV-19/Spain/RI-IBV-99010908/2020\|EPI_ISL_732754\| | ···················· | ···················· | ···················· | ···················· | NNNNNNNNNNNNNNNNNNNN | NNN················· | 1 |
| Y | 733 | hCoV-19/Spain/RI-IBV-99010909/2020\|EPI_ISL_732755\| | ···················· | ···················· | ···················· | ···················· | NNNNNNNNNNNNNNNNNNNN | NNNNNNNNNNNNNNN····· | 1 |
| Y | 734 | hCoV-19/Spain/RI-IBV-99010911/2020\|EPI_ISL_732756\| | ···················· | ···················· | ···················· | ···················· | NNNNNNNNNNNNNNNNN··· | ···················· | 1 |
| Y | 735 | hCoV-19/Spain/RI-IBV-99010912/2020\|EPI_ISL_732757\| | ···················· | ···················· | ···················· | ···················· | NNNNNNNNNNNNNNNNNNNN | NNNNNNNNNNNNNNNNNNNN | 1 |
| Y | 736 | hCoV-19/Spain/RI-IBV-99010918/2020\|EPI_ISL_732758\| | ···················· | ···················· | ···················· | ···················· | NNNNNNNNNNNNNNNNNNNN | NNNNNNNNNNNNNNNNNNNN | 1 |
| Y | 737 | hCoV-19/Spain/RI-IBV-99010919/2020\|EPI_ISL_732759\| | ···················· | ···················· | ···················· | ···················· | NNNNNNNNNNNNNNNNNNNN | NNNNNNNNNNNNNNNNNNNN | 1 |
| Y | 740 | hCoV-19/Spain/RI-IBV-99010925/2020\|EPI_ISL_732764\| | ···················· | ···················· | ···················· | ···················· | ··NNNNN············· | NNNNNNN············· | 1 |
| Y | 741 | hCoV-19/Spain/RI-IBV-99010926/2020\|EPI_ISL_732765\| | ···················· | N·····NNNNNNNNNNNNNN | ···················· | ···················· | ···················· | ···················· | 1 |
| Y | 742 | hCoV-19/Spain/RI-IBV-99010927/2020\|EPI_ISL_732766\| | ···················· | ···················· | ···················· | ···················· | NNNNNNNNNNNNNNNNNNNN | NNNNNNNNNNNNNNNNNNNN | 1 |
| Y | 743 | hCoV-19/Spain/RI-IBV-99010929/2020\|EPI_ISL_732767\| | ···················· | ···················· | ···················· | ···················· | NNNNNNNNNNNNNNNNNNNN | NNNNNNNNNNNNNNNNNNNN | 1 |
| Y | 745 | hCoV-19/Spain/RI-IBV-99010931/2020\|EPI_ISL_732769\| | ···················· | ···················· | ···················· | ···················· | NNNNNNN············· | ···················· | 1 |
| Y | 746 | hCoV-19/Spain/RI-IBV-99010932/2020\|EPI_ISL_732770\| | ···················· | ···················· | ···················· | ···················· | NNNNNNNNNNNNNNNNNNNN | NNNNNNNNNNNNNNNNNNNN | 1 |
| Y | 747 | hCoV-19/Spain/RI-IBV-99010934/2020\|EPI_ISL_732772\| | ···················· | ···················· | ···················· | ···················· | NNNNNNNNNNNNNNNNNNNN | NNNNNNNNNNNNNNNNNNNN | 1 |
| Y | 748 | hCoV-19/Spain/RI-IBV-99010936/2020\|EPI_ISL_732773\| | ···················· | ···················· | ··T················· | ···················· | NNNNNNNNNNNNNNNNNNNN | NNNNNNNNNNNNNNNNNNNN | 1 |
| Y | 749 | hCoV-19/Spain/RI-IBV-99010937/2020\|EPI_ISL_732774\| | ···················· | ···················· | ···················· | ···················· | ·NNNNNN······NNNNNNN | ···················· | 1 |
| Y | 750 | hCoV-19/Spain/RI-IBV-99010942/2020\|EPI_ISL_732777\| | ···················· | ···················· | ···················· | ···················· | NNNNNNNNNNNNNNNNNNNN | NNNNNNNNNNNNNNN····· | 1 |
| Y | 751 | hCoV-19/Spain/RI-IBV-99010947/2020\|EPI_ISL_732778\| | ···················· | ··········NNNNNNN··· | ···················· | ···················· | ···················· | ···················· | 1 |
| Y | 752 | hCoV-19/Spain/RI-IBV-99010948/2020\|EPI_ISL_732779\| | ···················· | NNNNNNNNNNNNNNNNNNNN | ···················· | ···················· | ·NNNNNN············· | ···················· | 1 |
| Y | 753 | hCoV-19/Spain/RI-IBV-99010949/2020\|EPI_ISL_732780\| | ···················· | ···················· | ···················· | ···················· | ··N··········NNNNNNN | ···················· | 1 |
| Y | 755 | hCoV-19/Spain/RI-IBV-99010956/2020\|EPI_ISL_732785\| | ···················· | ···················· | ···················· | ···················· | NNNNNNNNNNNNNNNNNNNN | NNNNNNNNNNNNNNNNNNNN | 1 |
| Y | 756 | hCoV-19/Spain/RI-IBV-99010958/2020\|EPI_ISL_732786\| | ···················· | ···················· | ···················· | ···················· | NNNNNNNNNNNNNNNNNNNN | NNNNNNNNNNNNNNNNNNNN | 1 |
| Y | 757 | hCoV-19/Spain/RI-IBV-99010959/2020\|EPI_ISL_732787\| | ···················· | ···················· | ···················· | ···················· | NNNNNNNNNNNNNNNNNNNN | NNNNNNNNNNNNNNNNNNNN | 1 |
| Y | 759 | hCoV-19/Spain/RI-IBV-99010961/2020\|EPI_ISL_732789\| | ···················· | ···················· | ···················· | NNNNNNNNNNNNNNNNNNNN | ···················· | ···················· | 1 |
| Y | 760 | hCoV-19/Spain/RI-IBV-99010964/2020\|EPI_ISL_732792\| | ···················· | NNNNNNNNNNNNNNNNNNNN | ···················· | ···················· | ·NNNNNNNNNNNNNNNN··· | ···················· | 1 |
| Y | 761 | hCoV-19/Spain/RI-IBV-99010965/2020\|EPI_ISL_732793\| | ···················· | ···················· | ···················· | ···················· | NNNNNNNNNNNNNNNNNNNN | NNNNNNNNNNNNNNNNNNNN | 1 |
| Y | 762 | hCoV-19/Spain/RI-IBV-99010967/2020\|EPI_ISL_732795\| | ···················· | ···················· | ···················· | ···················· | NNNNNNNNNNNNNNNNNNNN | NNNNNNNNNNNNNNNNNNNN | 1 |
| Y | 763 | hCoV-19/Spain/RI-IBV-99010968/2020\|EPI_ISL_732796\| | ···················· | ···················· | ···················· | ···················· | NNNNNNNNNNNNNNNNNNNN | ···················· | 1 |
| Y | 764 | hCoV-19/Spain/RI-IBV-99010969/2020\|EPI_ISL_732797\| | ···················· | ···················· | ···················· | ···················· | NNNNNNNNNNNNNNNNNNNN | NNNNNNNNNNNNNNNNNNNN | 1 |
| Y | 765 | hCoV-19/Spain/RI-IBV-99010970/2020\|EPI_ISL_732798\| | ···················· | ···················· | ···················· | ···················· | NNNNNNNNNNNNNNNNNNNN | NNNNNNNNNNNNNNNNNNNN | 1 |
| Y | 766 | hCoV-19/Spain/RI-IBV-99010971/2020\|EPI_ISL_732799\| | ···················· | ···················· | ···················· | ···················· | ··NNNNN············· | ···················· | 1 |
| Y | 785 | hCoV-19/Wales/PHWC-4925D3/2020\|EPI_ISL_725671\|2020 | ···················· | ···················· | ···················· | ···················· | ···················· | NNNNN······NNNNNNNNN | 1 |
| Y | 788 | hCoV-19/Wales/PHWC-49260D/2020\|EPI_ISL_725674\|2020 | ···················· | ···················· | ···················· | ···················· | NNNNNNNNNNNNNNNNNNNN | NNNNNNNNNNNNNNNNNNNN | 1 |
| Y | 790 | hCoV-19/Wales/PHWC-49262B/2020\|EPI_ISL_725676\|2020 | ···················· | ···················· | ···················· | ···················· | NNNNNNNNNNNNNNNNNNNN | NNNNNNNNNNNNNNNNNNNN | 1 |
| Y | 792 | hCoV-19/Wales/PHWC-492667/2020\|EPI_ISL_725679\|2020 | ···················· | ···················· | ···················· | ···················· | NNNNNNNNNNNNNNNNNNNN | NNNNNNNNNNNNNNNNNNNN | 1 |
| Y | 802 | hCoV-19/Wales/PHWC-497A84/2020\|EPI_ISL_726926\|2020 | ···················· | ···················· | ···················· | ···················· | NNNNNNNNNNNNNNNNNNNN | NN·················· | 1 |
| Y | 810 | hCoV-19/Wales/PHWC-497B09/2020\|EPI_ISL_726934\|2020 | ···················· | ···················· | ···················· | ···················· | NNNNNNNNNNNNNNNNNNNN | ···················· | 1 |
| Y | 823 | hCoV-19/Wales/PHWC-497C15/2020\|EPI_ISL_726949\|2020 | ···················· | ···················· | ···················· | ···················· | ···················· | NNNN················ | 1 |
| Y | 824 | hCoV-19/Wales/PHWC-497C33/2020\|EPI_ISL_726951\|2020 | ···················· | ···················· | ···················· | ···················· | NNNNNNNNNNNNNNNNN··· | ···················· | 1 |
| Y | 829 | hCoV-19/Wales/PHWC-497CAC/2020\|EPI_ISL_726958\|2020 | ···················· | ···················· | ···················· | ···················· | NNNNNNNNNNNNNNNNNNNN | NNNNNNNNNNNNNNNNNNNN | 1 |
| Y | 941 | hCoV-19/Belgium/rega-09221229/2020\|EPI_ISL_735024\| | ···················· | ···················· | ···················· | ···················· | NNNNNNNNNNNNNNNNNNNN | NNNNNNNNNNNNNNNNNNNN | 1 |
| Y | 973 | hCoV-19/Switzerland/VD-ETHZ-320525/2020\|EPI_ISL_73 | ···················· | ···················· | ···················· | ···················· | ····NNNNNNNNNNNNNNNN | ···················N | 1 |
| Y | 974 | hCoV-19/Switzerland/VD-ETHZ-320579/2020\|EPI_ISL_73 | ···················· | ···················· | ···················· | ···················· | ·······NNNNNNNNNNNNN | NNNNNNNNNNNNN······· | 1 |
| Y | 1000 | hCoV-19/Switzerland/AG-ETHZ-410117/2020\|EPI_ISL_73 | ···················· | ···················· | ···················· | ···················· | ·······NNNNNNNNNNNNN | ···················· | 1 |
| Y | 1003 | hCoV-19/Switzerland/AG-ETHZ-410213/2020\|EPI_ISL_73 | ···················· | ···················· | ···················· | ···················· | ·······NNNNNNNNNNNNN | ···················· | 1 |
| Y | 1010 | hCoV-19/Switzerland/BE-ETHZ-320201/2020\|EPI_ISL_73 | ···················· | ···················· | ···················· | ···················· | ····NNNNNNNNNNNNNNNN | ·······N····N······· | 1 |
| Y | 1012 | hCoV-19/Switzerland/BE-ETHZ-320580/2020\|EPI_ISL_73 | ···················· | ···················· | ···················· | ···················· | ····NNNNNNNNNN····NN | ···················· | 1 |
| Y | 1016 | hCoV-19/Switzerland/BE-ETHZ-320602/2020\|EPI_ISL_73 | ···················· | ···················· | ···················· | ···················· | ····NNNNNNNNNNNNNNNN | NNNNN·······NNN····· | 1 |
| Y | 1027 | hCoV-19/Wales/PHWC-4926A3/2020\|EPI_ISL_725683\|2020 | ···················· | ···················· | ···················· | ···················· | NNNNNNNNNNNN········ | NNNNNNNNNNNNNNN····· | 1 |
| Y | 1030 | hCoV-19/Wales/PHWC-492719/2020\|EPI_ISL_725690\|2020 | ···················· | ···················· | ···················· | ···················· | NNNNNNNNNNNNNNNNN··· | ···················· | 1 |
| Y | 1032 | hCoV-19/Wales/PHWC-492746/2020\|EPI_ISL_725693\|2020 | ···················· | ···················· | ···················· | ···················· | NNNNNNNNNNNNNNNNNNNN | NNNNNNNNNNNNNNNNNNNN | 1 |
| Y | 1050 | hCoV-19/Spain/RI-IBV-99010972/2020\|EPI_ISL_732800\| | ···················· | ···················· | ···················· | ···················· | NNNNNNNNNNNNNNNNNNNN | NNNNNNNNNNNNNNNNNNNN | 1 |
| Y | 1051 | hCoV-19/Spain/RI-IBV-99010973/2020\|EPI_ISL_732801\| | ···················· | ···················· | ···················· | ···················· | NNNNNNNNNNNNNNNNNNNN | NNNNNNNNNNNN········ | 1 |
| Y | 1052 | hCoV-19/Spain/RI-IBV-99010974/2020\|EPI_ISL_732802\| | ···················· | ···················· | ···················· | ···················· | NNNNNNNNNNNNNNNNNNNN | NNNNNNNNNNNNNNNNNNNN | 1 |
| Y | 1053 | hCoV-19/Spain/RI-IBV-99010975/2020\|EPI_ISL_732803\| | ···················· | ···················· | ···················· | ···················· | NNNNNNNNNNNNNNNNNNNN | NNNNNNNNNNNNNNNNNNNN | 1 |
| Y | 1054 | hCoV-19/Spain/RI-IBV-99010976/2020\|EPI_ISL_732804\| | ···················· | ···················· | ···················· | ···················· | NNNNNNNNNNNNNNNNNNNN | NNNNNNNNNNNNNNNNNNNN | 1 |
| Y | 1055 | hCoV-19/Spain/RI-IBV-99010977/2020\|EPI_ISL_732805\| | ···················· | ···················· | ···················· | ···················· | NNNNNNNNNNNNNNNNNNNN | NNNNNNNNNNNNNNNNNNNN | 1 |
| Y | 1056 | hCoV-19/Spain/RI-IBV-99010979/2020\|EPI_ISL_732806\| | ···················· | ···················· | ···················· | ···················· | NNNNNNNNNNNNNNNNNNNN | NNNNNNNNNNNNNNNNNNNN | 1 |
| Y | 1058 | hCoV-19/Spain/RI-IBV-99010981/2020\|EPI_ISL_732808\| | ···················· | ···················· | ···················· | ···················· | NNNNNNNNNNN········· | ···················· | 1 |
| Y | 1059 | hCoV-19/Spain/RI-IBV-99010986/2020\|EPI_ISL_732809\| | ···················· | ···················· | ···················· | ···················· | NNNNNNNNNNNNNNNNNNNN | NNNNNNNNNNNNNNNNNNNN | 1 |
| Y | 1060 | hCoV-19/Spain/RI-IBV-99010987/2020\|EPI_ISL_732810\| | ···················· | ···················· | ···················· | ···················· | NNNNNNNNNNNNNNNNNNNN | NNNNNNNNNNNNNNNNNNNN | 1 |
| Y | 1061 | hCoV-19/Spain/RI-IBV-99010988/2020\|EPI_ISL_732811\| | ···················· | ···················· | ···················· | ···················· | NNNNNNNNNNNNNNNNNNNN | NNNNNNNNNNNNNNNNNNNN | 1 |
| Y | 1062 | hCoV-19/Spain/RI-IBV-99010989/2020\|EPI_ISL_732812\| | ···················· | ···················· | ···················· | ···················· | NNNNNNNNNNNNNNNNNNNN | NNNNNNNNNNNNNNNNNNNN | 1 |
| Y | 1063 | hCoV-19/Spain/RI-IBV-99010990/2020\|EPI_ISL_732813\| | ···················· | ···················· | ···················· | ···················· | NNNNNNNNNNNNNNNNNNNN | NNNNNNNNNNNNNNNNNNNN | 1 |
| Y | 1064 | hCoV-19/Spain/RI-IBV-99010991/2020\|EPI_ISL_732814\| | ···················· | ···················· | ···················· | ···················· | NNNNNNNNNNNNNNNNNNNN | NNNNNNNNNNNNNNNNNNNN | 1 |
| Y | 1065 | hCoV-19/Spain/RI-IBV-99010992/2020\|EPI_ISL_732815\| | ···················· | ···················· | ···················· | ···················· | NNNNNNN············· | ···················· | 1 |
| Y | 1067 | hCoV-19/Spain/RI-IBV-99010994/2020\|EPI_ISL_732817\| | ···················· | ···················· | ···················· | ···················· | NNNNNNNNNNNNNNNNNNNN | NNNNNNNNNNNNNNNNNNNN | 1 |
| Y | 1068 | hCoV-19/Spain/RI-IBV-99010995/2020\|EPI_ISL_732818\| | ···················· | ···················· | ···················· | ···················· | NNNNNNNNNNNNNNNNNNNN | NNNNNNNNNNNNNNNNNNNN | 1 |
| Y | 1069 | hCoV-19/Spain/RI-IBV-99010996/2020\|EPI_ISL_732819\| | ···················· | ···················· | ···················· | ···················· | NNNNNNNNNNNNNNNNNNNN | NNNNNNNNNNNNNNNNNNNN | 1 |
| Y | 1169 | hCoV-19/Belgium/rega-10081257/2020\|EPI_ISL_735094\| | ···················· | NNNNNNNNNNNNNNNNNNNN | ···················· | ···················· | ···················· | ···················· | 1 |
| Y | 1200 | hCoV-19/Switzerland/BL-ETHZ-410136/2020\|EPI_ISL_73 | ···················· | ···················· | ···················· | ···················· | ·······NNNNNNNNNNNNN | ···················· | 1 |
| Y | 1204 | hCoV-19/Switzerland/BL-ETHZ-410182/2020\|EPI_ISL_73 | ···················· | ···················· | ···················· | ···················· | ·······NNNNNNNNNNNNN | ···················· | 1 |
| Y | 1216 | hCoV-19/Switzerland/BL-ETHZ-410962/2020\|EPI_ISL_73 | ···················· | ···················· | ···················· | ···················· | ·······NNNNNNNNNNNNN | NNNNNNNNNNNNNNNNNNNN | 1 |
| Y | 1285 | hCoV-19/Wales/PHWC-492834/2020\|EPI_ISL_725707\|2020 | ···················· | ···················· | ···················· | ···················· | NNNNNNNNNNNNNNNNNNNN | NNNNNNN············· | 1 |
| Y | 1288 | hCoV-19/Wales/PHWC-492861/2020\|EPI_ISL_725710\|2020 | ···················· | ···················· | ···················· | ···················· | NNNNNNNNNNNNNNNNNNNN | ···················· | 1 |
| Y | 1291 | hCoV-19/Wales/PHWC-4928BC/2020\|EPI_ISL_725714\|2020 | ···················· | ···················· | ···················· | ···················· | NNNNNNNNNNNNNNNNNNNN | NNNNNNNNNNNNNNN·NNNN | 1 |
| Y | 1306 | hCoV-19/Switzerland/VS-ETHZ-360332/2020\|EPI_ISL_72 | ···················· | ···················· | ···················· | ···················· | ·······NNNNNNNNNNNNN | NNNNNNNNNNNNNNNNNNNN | 1 |

| | | | | | | | | |
|---|---|---|---|---|---|---|---|---|
| Y | 1323 hCoV-19/Switzerland/BL-ETHZ-360277/2020\|EPI_ISL_72 | ···················· | ···················· | ···················· | ···················· | ·······NNNNNNNNNNNNN | NNNNNNNNNNNNNNNNNNNN | 1 |
| Y | 1326 hCoV-19/Switzerland/BL-ETHZ-360273/2020\|EPI_ISL_72 | ···················· | ···················· | ···················· | ···················· | ·······NNNNNNNNNNNNN | ···················· | 1 |
| Y | 1332 hCoV-19/Switzerland/LU-ETHZ-360260/2020\|EPI_ISL_72 | ···················· | ·A·················· | ···················· | ···················· | ·······NNNNNNNNNNNNN | ···················· | 1 |
| Y | 1337 hCoV-19/Switzerland/AG-ETHZ-360246/2020\|EPI_ISL_72 | ···················· | ···················· | ···················· | ···················· | ·······NNNNNNNNNNNNN | NNNNNNNNNNNNNNNNNNNN | 1 |
| Y | 1338 hCoV-19/Switzerland/AG-ETHZ-360245/2020\|EPI_ISL_72 | ···················· | ···················· | ···················· | ···················· | ·······NNNNNNNNNNNNN | ···················· | 1 |
| Y | 1545 hCoV-19/Wales/PHWC-492940/2020\|EPI_ISL_725723\|2020 | ···················· | ···················· | ···················· | ···················· | NNNNNNNNNNN········· | ···················· | 1 |
| Y | 1555 hCoV-19/Wales/PHWC-492A2F/2020\|EPI_ISL_725735\|2020 | ···················· | ···················· | ···················· | ···················· | ···················· | NNNNNNNNNNNNNNNNNNNN | 1 |
| Y | 1556 hCoV-19/Wales/PHWC-492A3E/2020\|EPI_ISL_725736\|2020 | ···················· | ···················· | ···················· | ···················· | NNNNNNNNNNNNNNNNNNNN | ···················· | 1 |
| Y | 1557 hCoV-19/Wales/PHWC-492A4D/2020\|EPI_ISL_725737\|2020 | ···················· | ···················· | ···················· | ···················· | NNNNNNNNNNNNNNNNNNNN | ···················· | 1 |
| Y | 1582 hCoV-19/Wales/PHWC-492C92/2020\|EPI_ISL_725772\|2020 | ···················· | ·····A·············· | ···················· | ···················· | NNNNNNNNNNNNNNNNNNNN | NNNNNNNNNNNNNNNNNNNN | 1 |
| Y | 1586 hCoV-19/Wales/PHWC-492CDE/2020\|EPI_ISL_725776\|2020 | ···················· | ···················· | ···················· | ···················· | NNNNNNNNNNNNNNNNNNNN | NNNNNNNNNNNNNNNNNNNN | 1 |
| Y | 1591 hCoV-19/Wales/PHWC-492D53/2020\|EPI_ISL_725782\|2020 | ···················· | ·····A·············· | ···················· | ···················· | NNNNNNNNNNNNNNNNNNNN | ···················· | 1 |
| Y | 1592 hCoV-19/Wales/PHWC-492D71/2020\|EPI_ISL_725783\|2020 | ···················· | ···················· | ···················· | ···················· | NNNNNNNNNNNNNNNNNNNN | NNNNNNNNNNNNNNNNNNNN | 1 |
| Y | 1600 hCoV-19/Wales/PHWC-492E05/2020\|EPI_ISL_725792\|2020 | ···················· | ···················· | ···················· | ···················· | NNNNNNNNNNNNNNNNNNNN | NNNNNNNNNNNNNNNNNNNN | 1 |
| Y | 1696 hCoV-19/Switzerland/BE-ETHZ-321087/2020\|EPI_ISL_73 | ···················· | ···················· | ···················· | ···················· | ··NNNNNNNNNNNNNNNNNN | NNNNNNNNNNNNNNNNNNNN | 1 |
| Y | 1710 hCoV-19/Switzerland/BE-ETHZ-410199/2020\|EPI_ISL_73 | ···················· | ···················· | ···················· | ···················· | ·······NNNNNNNNNNNNN | NNNNNNNNNNNNNNNNNNNN | 1 |
| Y | 1713 hCoV-19/Switzerland/BE-ETHZ-410204/2020\|EPI_ISL_73 | ···················· | ···················· | ···················· | ···················· | ·······NNNNNNNNNNNNN | NNNNNNNNNNNNNNNNNNNN | 1 |
| Y | 1727 hCoV-19/Switzerland/BE-ETHZ-410294/2020\|EPI_ISL_73 | ···················· | ···················· | ···················· | ···················· | ·······NNNNNNNNNNNNN | ···················· | 1 |
| Y | 1731 hCoV-19/Switzerland/BE-ETHZ-410563/2020\|EPI_ISL_73 | ···················· | ···················· | ···················· | ···················· | ·······NNNNNNNNNNNNN | ···················· | 1 |
| Y | 1775 hCoV-19/Switzerland/SH-ETHZ-410539/2020\|EPI_ISL_73 | ···················· | ·A·················· | ···················· | ···················· | ·······NNNNNNNNNNNNN | NNNNNNNNNNNNNNNNNNNN | 1 |
| Y | 1794 hCoV-19/USA/WI-UW-2249/2020\|EPI_ISL_728018\|2020-09 | ···················· | ···················· | ···················· | ···················· | NNNNNNNNNNNNNNNNNNNN | NNNNNNNNNNNNNNNNNNNN | 1 |
| Y | 1860 hCoV-19/Poland/19832/2020\|EPI_ISL_732921\|2020-11 | ···················· | NNNNNNNNNNNNNNNNNNNN | ···················· | ···················· | ···················· | ···················· | 1 |
| Y | 1915 hCoV-19/Belgium/rega-10191240/2020\|EPI_ISL_735166\| | ···················· | ···················· | ···················· | ···················· | NNNNNNNNNNNNNNNNNNNN | NNNNNNNNNNNNNNNNNNNN | 1 |
| Y | 1944 hCoV-19/Belgium/rega-10191289/2020\|EPI_ISL_735195\| | ···················· | ···················· | ···················· | ···················· | NNNNNNNNNNNNNNNNNNNN | NNNNNNNNNNNNNNNNNNNN | 1 |
| Y | 1978 hCoV-19/Switzerland/SO-ETHZ-410159/2020\|EPI_ISL_73 | ···················· | ···················· | ···················· | ···················· | ·······NNNNNNNNNNNNN | NNNNNNNNNNNNNNNNNNNN | 1 |
| Y | 2034 hCoV-19/USA/NC-CDC-2-3693565/2020\|EPI_ISL_747178\|2 | NNNNNNNNNNNNNNNNNNNN | NNNNNNNNNNNNNNNNNNNN | ···················· | ···················· | ···················· | ···················· | 1 |
| Y | 2059 hCoV-19/Wales/PHWC-49300E/2020\|EPI_ISL_725823\|2020 | ···················· | ···················· | ···················· | ···················· | NNNNNNNNNNNNNNNNNNNN | ···················· | 1 |
| Y | 2060 hCoV-19/Wales/PHWC-49301D/2020\|EPI_ISL_725824\|2020 | ···················· | ···················· | ···················· | ···················· | NNNNNNNNNNNNNNNNNN·· | ···················· | 1 |
| Y | 2068 hCoV-19/USA/WI-UW-2232/2020\|EPI_ISL_728002\|2020-10 | ···················· | ·A·················· | ···················· | ···················· | NNNNNNNNNNNNNNNNNNNN | NNNNNNNNNNNNNNNNNNNN | 1 |
| Y | 2074 hCoV-19/USA/WI-UW-2240/2020\|EPI_ISL_728009\|2020-11 | ···················· | ···················· | ···················· | ···················· | NNNNNNNNNNNNNNNNNNNN | NNNNNNNNNNNNNNNNNNNN | 1 |
| Y | 2077 hCoV-19/USA/WI-UW-2243/2020\|EPI_ISL_728012\|2020-09 | ···················· | ···················· | ···················· | ···················· | NNNNNNNNNNNNNNNNNNNN | NNNNNNNNNNNNNNNNNNNN | 1 |
| Y | 2082 hCoV-19/USA/WI-UW-2248/2020\|EPI_ISL_728017\|2020-10 | ···················· | ·A·················· | ···················· | ···················· | NNNNNNNNNNNNNNNNNNNN | NNNNNNNNNNNNNNNNNNNN | 1 |
| Y | 2083 hCoV-19/USA/WI-UW-2251/2020\|EPI_ISL_728020\|2020-09 | ···················· | ···················· | ···················· | ···················· | NNNNNNNNNNNNNNNNNNNN | NNNNNNNNNNNNNNNNNNNN | 1 |
| Y | 2087 hCoV-19/USA/WI-UW-2255/2020\|EPI_ISL_728024\|2020-09 | ···················· | ·A·················· | ···················· | ···················· | NNNNNNNNNNNNNNNNNNNN | NNNNNNNNNNNNNNNNNNNN | 1 |
| Y | 2098 hCoV-19/USA/WI-UW-2267/2020\|EPI_ISL_728036\|2020-10 | ···················· | ···················· | ···················· | ···················· | NNNNNNNNNNNNNNNNNNNN | NNNNNNNNNNNNNNNNNNNN | 1 |
| Y | 2099 hCoV-19/USA/WI-UW-2268/2020\|EPI_ISL_728037\|2020-10 | ···················· | ···················· | ···················· | ···················· | NNNNNNNNNNNNNNNNNNNN | NNNNNNNNNNNNNNNNNNNN | 1 |
| Y | 2100 hCoV-19/USA/WI-UW-2269/2020\|EPI_ISL_728038\|2020-11 | ···················· | ···················· | ···················· | ···················· | NNNNNNNNNNNNNNNNNNNN | NNNNNNNNNNNNNNNNNNNN | 1 |
| Y | 2102 hCoV-19/USA/WI-UW-2271/2020\|EPI_ISL_728040\|2020-11 | ···················· | ···················· | ···················· | ···················· | NNNNNNNNNNNNNNNNNNNN | NNNNNNNNNNNNNNNNNNNN | 1 |
| Y | 2104 hCoV-19/USA/WI-UW-2275/2020\|EPI_ISL_728042\|2020-11 | ···················· | ···················· | ···················· | ···················· | NNNNNNNNNNNNNNNNNNNN | NNNNNNNNNNNNNNNNNNNN | 1 |
| Y | 2105 hCoV-19/USA/WI-UW-2276/2020\|EPI_ISL_728043\|2020-11 | ···················· | ···················· | ···················· | ···················· | NNNNNNNNNNNNNNNNNNNN | NNNNNNNNNNNNNNNNNNNN | 1 |
| Y | 2107 hCoV-19/USA/WI-UW-2278/2020\|EPI_ISL_728045\|2020-11 | ···················· | ···················· | ···················· | ···················· | NNNNNNNNNNNNNNNNNNNN | NNNNNNNNNNNNNNNNNNNN | 1 |
| Y | 2109 hCoV-19/USA/WI-UW-2280/2020\|EPI_ISL_728047\|2020-11 | ···················· | ···················· | ···················· | ···················· | NNNNNNNNNNNNNNNNNNNN | NNNNNNNNNNNNNNNNNNNN | 1 |
| Y | 2110 hCoV-19/USA/WI-UW-2281/2020\|EPI_ISL_728048\|2020-11 | ···················· | ···················· | ···················· | ···················· | NNNNNNNNNNNNNNNNNNNN | NNNNNNNNNNNNNNNNNNNN | 1 |
| Y | 2111 hCoV-19/USA/WI-UW-2282/2020\|EPI_ISL_728049\|2020-11 | ···················· | ···················· | ···················· | ···················· | NNNNNNNNNNNNNNNNNNNN | NNNNNNNNNNNNNNNNNNNN | 1 |
| Y | 2112 hCoV-19/USA/WI-UW-2283/2020\|EPI_ISL_728050\|2020-10 | ···················· | ···················· | ···················· | ···················· | NNNNNNNNNNNNNNNNNNNN | NNNNNNNNNNNNNNNNNNNN | 1 |
| Y | 2113 hCoV-19/USA/WI-UW-2285/2020\|EPI_ISL_728051\|2020-10 | ···················· | ··C················· | ···················· | ···················· | NNNNNNNNNNNNNNNNNNNN | NNNNNNNNNNNNNNNNNNNN | 1 |
| Y | 2114 hCoV-19/USA/WI-UW-2286/2020\|EPI_ISL_728052\|2020-10 | ···················· | ···················· | ···················· | ···················· | NNNNNNNNNNNNNNNNNNNN | NNNNNNNNNNNNNNNNNNNN | 1 |
| Y | 2115 hCoV-19/USA/WI-UW-2287/2020\|EPI_ISL_728053\|2020-11 | ···················· | ···················· | ···················· | ···················· | NNNNNNNNNNNNNNNNNNNN | NNNNNNNNNNNNNNNNNNNN | 1 |
| Y | 2116 hCoV-19/USA/WI-UW-2288/2020\|EPI_ISL_728054\|2020-11 | ···················· | ···················· | ···················· | ···················· | NNNNNNNNNNNNNNNNNNNN | NNNNNNNNNNNNNNNNNNNN | 1 |
| Y | 2118 hCoV-19/USA/WI-UW-2290/2020\|EPI_ISL_728056\|2020-11 | ···················· | ···················· | ···················· | ···················· | NNNNNNNNNNNNNNNNNNNN | NNNNNNNNNNNNNNNNNNNN | 1 |
| Y | 2119 hCoV-19/USA/WI-UW-2291/2020\|EPI_ISL_728057\|2020-11 | ···················· | ···················· | ···················· | ···················· | NNNNNNNNNNNNNNNNNNNN | NNNNNNNNNNNNNNNNNNNN | 1 |
| Y | 2120 hCoV-19/USA/WI-UW-2292/2020\|EPI_ISL_728058\|2020-11 | ···················· | ···················· | ···················· | ···················· | NNNNNNNNNNNNNNNNNNNN | NNNNNNNNNNNNNNNNNNNN | 1 |
| Y | 2121 hCoV-19/USA/WI-UW-2293/2020\|EPI_ISL_728059\|2020-11 | ···················· | ···················· | ···················· | ···················· | NNNNNNNNNNNNNNNNNNNN | NNNNNNNNNNNNNNNNNNNN | 1 |
| Y | 2122 hCoV-19/USA/WI-UW-2294/2020\|EPI_ISL_728060\|2020-11 | ···················· | ···················· | ···················· | ···················· | NNNNNNNNNNNNNNNNNNNN | NNNNNNNNNNNNNNNNNNNN | 1 |
| Y | 2123 hCoV-19/USA/WI-UW-2295/2020\|EPI_ISL_728061\|2020-11 | ···················· | ···················· | ···················· | ···················· | NNNNNNNNNNNNNNNNNNNN | NNNNNNNNNNNNNNNNNNNN | 1 |
| Y | 2124 hCoV-19/USA/WI-UW-2296/2020\|EPI_ISL_728062\|2020-11 | ···················· | ···················· | ···················· | ···················· | NNNNNNNNNNNNNNNNNNNN | NNNNNNNNNNNNNNNNNNNN | 1 |
| Y | 2125 hCoV-19/USA/WI-UW-2297/2020\|EPI_ISL_728063\|2020-11 | ···················· | ···················· | ···················· | ···················· | NNNNNNNNNNNNNNNNNNNN | NNNNNNNNNNNNNNNNNNNN | 1 |
| Y | 2126 hCoV-19/USA/WI-UW-2298/2020\|EPI_ISL_728064\|2020-11 | ···················· | ···················· | ···················· | ···················· | NNNNNNNNNNNNNNNNNNNN | NNNNNNNNNNNNNNNNNNNN | 1 |
| Y | 2127 hCoV-19/USA/WI-UW-2299/2020\|EPI_ISL_728065\|2020-11 | ···················· | ···················· | ···················· | ···················· | NNNNNNNNNNNNNNNNNNNN | NNNNNNNNNNNNNNNNNNNN | 1 |
| Y | 2128 hCoV-19/USA/WI-UW-2300/2020\|EPI_ISL_728066\|2020-11 | ···················· | ···················· | ···················· | ···················· | NNNNNNNNNNNNNNNNNNNN | NNNNNNNNNNNNNNNNNNNN | 1 |
| Y | 2129 hCoV-19/USA/WI-UW-2301/2020\|EPI_ISL_728067\|2020-11 | ···················· | ···················· | ···················· | ···················· | NNNNNNNNNNNNNNNNNNNN | NNNNNNNNNNNNNNNNNNNN | 1 |
| Y | 2130 hCoV-19/USA/WI-UW-2302/2020\|EPI_ISL_728068\|2020-11 | ···················· | ···················· | ···················· | ···················· | NNNNNNNNNNNNNNNNNNNN | NNNNNNNNNNNNNNNNNNNN | 1 |
| Y | 2131 hCoV-19/USA/WI-UW-2303/2020\|EPI_ISL_728069\|2020-11 | ···················· | ···················· | ···················· | ···················· | NNNNNNNNNNNNNNNNNNNN | NNNNNNNNNNNNNNNNNNNN | 1 |
| Y | 2138 hCoV-19/USA/WI-UW-2330/2020\|EPI_ISL_728096\|2020-11 | ···················· | ···················· | ···················· | ···················· | NNNNNNNNNNNNNNNNNNNN | NNNNNNNNNNNNNNNNNNNN | 1 |
| Y | 2279 hCoV-19/Russia/KR-RII-12753S/2020\|EPI_ISL_732976\|2 | ···················· | ···················· | ···················· | ···················· | NNNNNNNNNNNNNNNNNNNN | ···················· | 1 |
| Y | 2280 hCoV-19/Russia/KR-RII-12236S/2020\|EPI_ISL_732979\|2 | ···················· | ···················· | ···················· | ···················· | NNNNNNNNNNNNNNNNNNNN | ···················· | 1 |
| Y | 2296 hCoV-19/Switzerland/SO-ETHZ-410842/2020\|EPI_ISL_73 | ···················· | ···················· | ···················· | ···················· | ·······NNNNNNNNNNNNN | ···················· | 1 |
| Y | 2325 hCoV-19/Wales/PHWC-492EAB/2020\|EPI_ISL_725802\|2020 | ···················· | ···················· | ···················· | ···················· | NNNNNNNNNNNNNNNNNNNN | ···················· | 1 |
| Y | 2326 hCoV-19/Wales/PHWC-492EBA/2020\|EPI_ISL_725803\|2020 | ···················· | ···················· | ···················· | ···················· | NNNNNNNNNNNNNNNNNNNN | ···················· | 1 |
| Y | 2327 hCoV-19/Wales/PHWC-492ED8/2020\|EPI_ISL_725804\|2020 | ···················· | ···················· | ···················· | ···················· | NNNNNNNNNNNNNNNNNNNN | ···················· | 1 |
| Y | 2330 hCoV-19/Wales/PHWC-492F11/2020\|EPI_ISL_725808\|2020 | ···················· | ···················· | ···················· | ···················· | NNNNNNNNNNNNNNNNNNNN | ···················· | 1 |
| Y | 2338 hCoV-19/Wales/PHWC-493068/2020\|EPI_ISL_725829\|2020 | ···················· | ···················· | ···················· | ···················· | NNNNNNNNNNNNNNNNNNNN | ········NNNNNN······ | 1 |
| Y | 2352 hCoV-19/Wales/PHWC-493192/2020\|EPI_ISL_725848\|2020 | ···················· | ···················· | ···················· | ···················· | NNNNNNNNNNNNNNNNNNNN | NNNNNNNNNNNNNNNNNNNN | 1 |
| Y | 2360 hCoV-19/Wales/PHWC-493226/2020\|EPI_ISL_725857\|2020 | ···················· | ···················· | ···················· | ···················· | NNNNNNNNNNNNNNNNNNNN | NNNNNNNNNNNNNNNNNNNN | 1 |
| Y | 2361 hCoV-19/Wales/PHWC-493262/2020\|EPI_ISL_725860\|2020 | ···················· | ···················· | ···················· | ···················· | NNNNNNNNNNNNNNNNNNNN | NNNNNNNNNNNNNNNNNNNN | 1 |
| Y | 2363 hCoV-19/Wales/PHWC-4932AE/2020\|EPI_ISL_725863\|2020 | ···················· | ···················· | ···················· | ···················· | NNNNNNNNNNNNNNNNNNNN | ···················· | 1 |
| Y | 2365 hCoV-19/Wales/PHWC-4932CC/2020\|EPI_ISL_725865\|2020 | ···················· | ···················· | ···················· | ···················· | ··NNNNNNNNNNNNNNNNNN | ···················· | 1 |
| Y | 2368 hCoV-19/Wales/PHWC-493305/2020\|EPI_ISL_725868\|2020 | ···················· | ···················· | ···················· | ···················· | NNNNNNNNNNN········· | ···················· | 1 |
| Y | 2371 hCoV-19/Wales/PHWC-493341/2020\|EPI_ISL_725871\|2020 | ···················· | ···················· | ···················· | ···················· | NNNNNNNNNNNNNNNNNNNN | ···················· | 1 |
| Y | 2373 hCoV-19/Wales/PHWC-49337E/2020\|EPI_ISL_725874\|2020 | ···················· | ···················· | ···················· | ···················· | NNNNNNNNNNNNNNNNNNNN | NNNNNNNNNNNNNNNNNNNN | 1 |

| | | | | | | | | |
|---|---|---|---|---|---|---|---|---|
| Y | 2376 hCoV-19/Wales/PHWC-4933AB/2020\|EPI_ISL_725877\|2020 | ···················· | ···················· | ···················· | ···················· | NNNNNNNNNNNNNNNNNNNN | NNNNN··············· | 1 |
| Y | 2378 hCoV-19/Wales/PHWC-4933C9/2020\|EPI_ISL_725879\|2020 | ···················· | ···················· | ···················· | ···················· | NNNNNNNNNNNNNNNNNNNN | NNNNNNNNNNNNNNN·NNNN | 1 |
| Y | 2382 hCoV-19/Wales/PHWC-493420/2020\|EPI_ISL_725885\|2020 | ···················· | ···················· | ···················· | ···················· | NNNNNNNNNNNNNNNNNNNN | NNNNNNNNNNNNNNNNNNNN | 1 |
| Y | 2387 hCoV-19/Wales/PHWC-4934C6/2020\|EPI_ISL_725895\|2020 | ···················· | ···················· | ···················· | ···················· | ···················· | NNNNNNNNNNNNNNNNNNNN | 1 |
| Y | 2391 hCoV-19/USA/WI-UW-2304/2020\|EPI_ISL_728070\|2020-11 | ···················· | ···················· | ···················· | ···················· | NNNNNNNNNNNNNNNNNNNN | NNNNNNNNNNNNNNNNNNNN | 1 |
| Y | 2393 hCoV-19/USA/WI-UW-2306/2020\|EPI_ISL_728072\|2020-11 | ···················· | ···················· | ···················· | ···················· | NNNNNNNNNNNNNNNNNNNN | NNNNNNNNNNNNNNNNNNNN | 1 |
| Y | 2394 hCoV-19/USA/WI-UW-2307/2020\|EPI_ISL_728073\|2020-11 | ···················· | ·A·················· | ···················· | ···················· | NNNNNNNNNNNNNNNNNNNN | NNNNNNNNNNNNNNNNNNNN | 1 |
| Y | 2395 hCoV-19/USA/WI-UW-2308/2020\|EPI_ISL_728074\|2020-11 | ···················· | ···················· | ···················· | ···················· | NNNNNNNNNNNNNNNNNNNN | NNNNNNNNNNNNNNNNNNNN | 1 |
| Y | 2396 hCoV-19/USA/WI-UW-2309/2020\|EPI_ISL_728075\|2020-11 | ···················· | ···················· | ···················· | ···················· | NNNNNNNNNNNNNNNNNNNN | NNNNNNNNNNNNNNNNNNNN | 1 |
| Y | 2397 hCoV-19/USA/WI-UW-2310/2020\|EPI_ISL_728076\|2020-10 | ···················· | ···················· | ···················· | ···················· | NNNNNNNNNNNNNNNNNNNN | NNNNNNNNNNNNNNNNNNNN | 1 |
| Y | 2398 hCoV-19/USA/WI-UW-2311/2020\|EPI_ISL_728077\|2020-10 | ···················· | ···················· | ···················· | ···················· | NNNNNNNNNNNNNNNNNNNN | NNNNNNNNNNNNNNNNNNNN | 1 |
| Y | 2399 hCoV-19/USA/WI-UW-2312/2020\|EPI_ISL_728078\|2020-11 | ···················· | ···················· | ···················· | ···················· | NNNNNNNNNNNNNNNNNNNN | NNNNNNNNNNNNNNNNNNNN | 1 |
| Y | 2400 hCoV-19/USA/WI-UW-2313/2020\|EPI_ISL_728079\|2020-11 | ···················· | ···················· | ···················· | ···················· | NNNNNNNNNNNNNNNNNNNN | NNNNNNNNNNNNNNNNNNNN | 1 |
| Y | 2402 hCoV-19/USA/WI-UW-2315/2020\|EPI_ISL_728081\|2020-11 | ···················· | ···················· | ···················· | ···················· | NNNNNNNNNNNNNNNNNNNN | NNNNNNNNNNNNNNNNNNNN | 1 |
| Y | 2404 hCoV-19/USA/WI-UW-2317/2020\|EPI_ISL_728083\|2020-11 | ···················· | ···················· | ···················· | ···················· | NNNNNNNNNNNNNNNNNNNN | NNNNNNNNNNNNNNNNNNNN | 1 |
| Y | 2405 hCoV-19/USA/WI-UW-2319/2020\|EPI_ISL_728085\|2020-11 | ···················· | ···················· | ···················· | ···················· | NNNNNNNNNNNNNNNNNNNN | NNNNNNNNNNNNNNNNNNNN | 1 |
| Y | 2486 hCoV-19/Switzerland/SO-ETHZ-410831/2020\|EPI_ISL_73 | ···················· | ···················· | ···················· | ···················· | ·······NNNNNNNNNNNNN | ···················· | 1 |
| Y | 2498 hCoV-19/Switzerland/VS-ETHZ-320200/2020\|EPI_ISL_73 | ···················· | ···················· | ···················· | ···················· | ···················· | NNNNNNNNNNNNNNNNNNNN | 1 |
| Y | 2499 hCoV-19/Switzerland/VS-ETHZ-320202/2020\|EPI_ISL_73 | ···················· | ···················· | ···················· | ···················· | ·······NNNNNNNNNNNNN | ···················· | 1 |
| Y | 2501 hCoV-19/Switzerland/VS-ETHZ-320206/2020\|EPI_ISL_73 | ···················· | ···················· | ···················· | ···················· | ······NNNNNNNNNNNNNN | NNNNNNNNNNNNNNNNNNNN | 1 |
| Y | 2507 hCoV-19/Switzerland/VS-ETHZ-320585/2020\|EPI_ISL_73 | ···················· | ···················· | ···················· | ···················· | ··NNNNNNNNNNNNNNNNNN | NNNNNNNNNNNNNNNNNNNN | 1 |
| Y | 2546 hCoV-19/Switzerland/VD-ETHZ-320405/2020\|EPI_ISL_73 | ···················· | ···················· | ···················· | ···················· | ··NNNNNNNNNNNNNNNNNN | NNNNNNNNNNNNNNNNNNNN | 1 |
| Y | 2557 hCoV-19/Switzerland/VD-ETHZ-320523/2020\|EPI_ISL_73 | ···················· | ···················· | ···················· | ···················· | ··NNNNNNNNNNNNNNNNNN | NNNNNNNNNNNNNNNNNNNN | 1 |
| Y | 2593 hCoV-19/Wales/PHWC-4934A8/2020\|EPI_ISL_725893\|2020 | ···················· | ···················· | ···················· | ···················· | NNNNNNNNNNN········· | ···················· | 1 |
| Y | 2596 hCoV-19/Wales/PHWC-4955FE/2020\|EPI_ISL_726371\|2020 | ···················· | ···················· | ···················· | ···················· | NNNNNNNNNNNNNNNNNNNN | NNNNNNNNNNNNNNNNNNNN | 1 |
| Y | 2597 hCoV-19/Wales/PHWC-49560A/2020\|EPI_ISL_726372\|2020 | ···················· | ···················· | ···················· | ···················· | NNNNNNNNNNNNNNNNNNNN | ···················· | 1 |
| Y | 2603 hCoV-19/Wales/PHWC-4956BF/2020\|EPI_ISL_726381\|2020 | ···················· | ···················· | ···················· | ···················· | NNNNNNNNNNNNNNNN···· | ···················· | 1 |
| Y | 2604 hCoV-19/Wales/PHWC-4956CE/2020\|EPI_ISL_726382\|2020 | ···················· | ······A············· | ···················· | ···················· | NNNNNNNNNNNNNNNNNNNN | NNNNNNNNNNN·····NNNN | 1 |
| Y | 2605 hCoV-19/Wales/PHWC-495707/2020\|EPI_ISL_726386\|2020 | ···················· | ···················· | ···················· | ···················· | NNNNNNNNNNNNNNNNNNNN | ···················· | 1 |
| Y | 2606 hCoV-19/Wales/PHWC-495716/2020\|EPI_ISL_726387\|2020 | ···················· | ···················· | ···················· | ···················· | NNNNNNNNNNNNNNNNNNNN | NNNNNNNNNNNNNNNNNNNN | 1 |
| Y | 2612 hCoV-19/Wales/PHWC-49579E/2020\|EPI_ISL_726394\|2020 | ···················· | ···················· | ···················· | ···················· | NNNNNNNNNNNNNNNNNNNN | NNNNNNNNNNNNNNNNNNNN | 1 |
| Y | 2615 hCoV-19/Wales/PHWC-4957CB/2020\|EPI_ISL_726397\|2020 | ···················· | ···················· | ···················· | ···················· | NNNNNNNNNNNNNNNNNNNN | NNNNNNNNNNNNNNNNNNNN | 1 |
| Y | 2652 hCoV-19/Belgium/rega-10191305/2020\|EPI_ISL_735208\| | ···················· | ···················· | ···················· | ···················· | NNNNNNNNNNNNNNNNNNNN | NNNNNNNNNNNNNNNNNNNN | 1 |
| Y | 2825 hCoV-19/Portugal/PT1977/2020\|EPI_ISL_732200\|2020-1 | ···················· | ···················· | ···················· | ···················· | NNNNNNNNNN··N······· | NNNNNNNNNNN·NNNNN··· | 1 |
| Y | 2875 hCoV-19/Portugal/PT2063/2020\|EPI_ISL_732277\|2020-1 | ···················· | ···················· | ···················· | ···················· | ·NNNNNNNNNNNNNNNN·NN | ···················· | 1 |
| Y | 2913 hCoV-19/Thailand/SamutSakhon-CONI-0334/2020\|EPI_IS | ···················· | ·A·················· | ···················· | ···················· | NNNNNNNNNNNNNNNNNNNN | NNNNNNNNNNNNNNNNNNNN | 1 |
| Y | 2914 hCoV-19/Thailand/SamutSakhon-CONI-0335/2020\|EPI_IS | ···················· | ·A·················· | ···················· | ···················· | NNNNNNNNNNNNNNNNNNNN | NNNNNNNNNNNNNNNNNNNN | 1 |
| Y | 2915 hCoV-19/Thailand/SamutSakhon-CONI-0336/2020\|EPI_IS | ···················· | ·A·················· | ···················· | ···················· | NNNNNNNNNNNNNNNNNNNN | NNNNNNNNNNNNNNNNNNNN | 1 |
| Y | 2916 hCoV-19/Thailand/SamutSakhon-CONI-0337/2020\|EPI_IS | ···················· | ·A·················· | ···················· | ···················· | NNNNNNNNNNNNNNNNNNNN | NNNNNNNNNNNNNNNNNNNN | 1 |
| Y | 2917 hCoV-19/Thailand/SamutSakhon-CONI-0338/2020\|EPI_IS | ···················· | ·A·················· | ···················· | ···················· | NNNNNNNNNNNNNNNNNNNN | NNNNNNNNNNNNNNNNNNNN | 1 |
| Y | 2918 hCoV-19/Thailand/SamutSakhon-CONI-0339/2020\|EPI_IS | ···················· | ·A·················· | ···················· | ···················· | NNNNNNNNNNNNNNNNNNNN | NNNNNNNNNNNNNNNNNNNN | 1 |
| Y | 2919 hCoV-19/Thailand/SamutSakhon-CONI-0340/2020\|EPI_IS | ···················· | ·A·················· | ···················· | ···················· | NNNNNNNNNNNNNNNNNNNN | NNNNNNNNNNNNNNNNNNNN | 1 |
| Y | 2920 hCoV-19/Thailand/SamutSakhon-CONI-0341/2020\|EPI_IS | ···················· | ·A·················· | ···················· | ···················· | NNNNNNNNNNNNNNNNNNNN | NNNNNNNNNNNNNNNNNNNN | 1 |
| Y | 2921 hCoV-19/Thailand/SamutSakhon-CONI-0342/2020\|EPI_IS | ···················· | ·A·················· | ···················· | ···················· | NNNNNNNNNNNNNNNNNNNN | NNNNNNNNNNNNNNNNNNNN | 1 |
| Y | 2922 hCoV-19/Thailand/SamutSakhon-CONI-0343/2020\|EPI_IS | ···················· | ·A·················· | ···················· | ···················· | NNNNNNNNNNNNNNNNNNNN | NNNNNNNNNNNNNNNNNNNN | 1 |
| Y | 2923 hCoV-19/Thailand/SamutSakhon-CONI-0344/2020\|EPI_IS | ···················· | ·A·················· | ···················· | ···················· | NNNNNNNNNNNNNNNNNNNN | NNNNNNNNNNNNNNNNNNNN | 1 |
| Y | 2924 hCoV-19/Thailand/SamutSakhon-CONI-0345/2020\|EPI_IS | ···················· | ·A·················· | ···················· | ···················· | NNNNNNNNNNNNNNNNNNNN | NNNNNNNNNNNNNNNNNNNN | 1 |
| Y | 2925 hCoV-19/Thailand/SamutSakhon-CONI-0346/2020\|EPI_IS | ···················· | ·A·················· | ···················· | ···················· | NNNNNNNNNNNNNNNNNNNN | NNNNNNNNNNNNNNNNNNNN | 1 |
| Y | 2926 hCoV-19/Thailand/SamutSakhon-CONI-0347/2020\|EPI_IS | ···················· | ·A·················· | ···················· | ···················· | NNNNNNNNNNNNNNNNNNNN | NNNNNNNNNNNNNNNNNNNN | 1 |
| Y | 2927 hCoV-19/Thailand/SamutSakhon-CONI-0348/2020\|EPI_IS | ···················· | ·A·················· | ···················· | ···················· | NNNNNNNNNNNNNNNNNNNN | NNNNNNNNNNNNNNNNNNNN | 1 |
| Y | 2928 hCoV-19/Thailand/SamutSakhon-CONI-0349/2020\|EPI_IS | ···················· | ·A·················· | ···················· | ···················· | NNNNNNNNNNNNNNNNNNNN | NNNNNNNNNNNNNNNNNNNN | 1 |
| Y | 2929 hCoV-19/Thailand/SamutSakhon-CONI-0350/2020\|EPI_IS | ···················· | ·A·················· | ···················· | ···················· | NNNNNNNNNNNNNNNNNNNN | NNNNNNNNNNNNNNNNNNNN | 1 |
| Y | 2930 hCoV-19/Thailand/SamutSakhon-CONI-0351/2020\|EPI_IS | ···················· | ·A·················· | ···················· | ···················· | NNNNNNNNNNNNNNNNNNNN | NNNNNNNNNNNNNNNNNNNN | 1 |
| Y | 2931 hCoV-19/Thailand/SamutSakhon-CONI-0352/2020\|EPI_IS | ···················· | ·A·················· | ···················· | ···················· | NNNNNNNNNNNNNNNNNNNN | NNNNNNNNNNNNNNNNNNNN | 1 |
| Y | 2932 hCoV-19/Thailand/SamutSakhon-CONI-0354/2020\|EPI_IS | ···················· | ·A·················· | ···················· | ···················· | NNNNNNNNNNNNNNNNNNNN | NNNNNNNNNNNNNNNNNNNN | 1 |
| Y | 2933 hCoV-19/Thailand/SamutSakhon-CONI-0355/2020\|EPI_IS | ···················· | ·A·················· | ···················· | ···················· | NNNNNNNNNNNNNNNNNNNN | NNNNNNNNNNNNNNNNNNNN | 1 |
| Y | 2934 hCoV-19/Thailand/SamutSakhon-CONI-0356/2020\|EPI_IS | ···················· | ·A·················· | ···················· | ···················· | NNNNNNNNNNNNNNNNNNNN | NNNNNNNNNNNNNNNNNNNN | 1 |
| Y | 2935 hCoV-19/Thailand/SamutSakhon-CONI-0359/2020\|EPI_IS | ···················· | ·A·················· | ···················· | ···················· | NNNNNNNNNNNNNNNNNNNN | NNNNNNNNNNNNNNNNNNNN | 1 |
| Y | 2936 hCoV-19/Thailand/SamutSakhon-CONI-0360/2020\|EPI_IS | ···················· | ·A·················· | ···················· | ···················· | NNNNNNNNNNNNNNNNNNNN | NNNNNNNNNNNNNNNNNNNN | 1 |
| Y | 2937 hCoV-19/Thailand/SamutSakhon-CONI-0361/2020\|EPI_IS | ···················· | ·A·················· | ···················· | ···················· | NNNNNNNNNNNNNNNNNNNN | NNNNNNNNNNNNNNNNNNNN | 1 |
| Y | 2938 hCoV-19/Thailand/SamutSakhon-CONI-0362/2020\|EPI_IS | ···················· | ·A·················· | ···················· | ···················· | NNNNNNNNNNNNNNNNNNNN | NNNNNNNNNNNNNNNNNNNN | 1 |
| Y | 2939 hCoV-19/Thailand/SamutSakhon-CONI-0363/2020\|EPI_IS | ···················· | ·A·················· | ···················· | ···················· | NNNNNNNNNNNNNNNNNNNN | NNNNNNNNNNNNNNNNNNNN | 1 |
| Y | 2940 hCoV-19/Thailand/SamutSakhon-CONI-0364/2020\|EPI_IS | ···················· | ·A·················· | ···················· | ···················· | NNNNNNNNNNNNNNNNNNNN | NNNNNNNNNNNNNNNNNNNN | 1 |
| Y | 2941 hCoV-19/Thailand/SamutSakhon-CONI-0365/2020\|EPI_IS | ···················· | ·A·················· | ···················· | ···················· | NNNNNNNNNNNNNNNNNNNN | NNNNNNNNNNNNNNNNNNNN | 1 |
| Y | 2942 hCoV-19/Thailand/SamutSakhon-CONI-0366/2020\|EPI_IS | ···················· | ·A·················· | ···················· | ···················· | NNNNNNNNNNNNNNNNNNNN | NNNNNNNNNNNNNNNNNNNN | 1 |
| Y | 2943 hCoV-19/Thailand/SamutSakhon-CONI-0367/2020\|EPI_IS | ···················· | ·A·················· | ···················· | ···················· | NNNNNNNNNNNNNNNNNNNN | NNNNNNNNNNNNNNNNNNNN | 1 |
| Y | 2944 hCoV-19/Thailand/SamutSakhon-CONI-0368/2020\|EPI_IS | ···················· | ·A·················· | ···················· | ···················· | NNNNNNNNNNNNNNNNNNNN | NNNNNNNNNNNNNNNNNNNN | 1 |
| Y | 2945 hCoV-19/Thailand/SamutSakhon-CONI-0369/2020\|EPI_IS | ···················· | ·A·················· | ···················· | ···················· | NNNNNNNNNNNNNNNNNNNN | NNNNNNNNNNNNNNNNNNNN | 1 |
| Y | 2946 hCoV-19/Thailand/SamutSakhon-CONI-0371/2020\|EPI_IS | ···················· | ·A·················· | ···················· | ···················· | NNNNNNNNNNNNNNNNNNNN | NNNNNNNNNNNNNNNNNNNN | 1 |
| Y | 2947 hCoV-19/Thailand/SamutSakhon-CONI-0373/2020\|EPI_IS | ···················· | ·A·················· | ···················· | ···················· | NNNNNNNNNNNNNNNNNNNN | NNNNNNNNNNNNNNNNNNNN | 1 |
| Y | 3113 hCoV-19/Wales/PHWC-4957F8/2020\|EPI_ISL_726400\|2020 | ···················· | ······A············· | ···················· | ···················· | NNNNNNNNNNNNNNNNNNNN | NNNNNNNNNNNNNNNNNNNN | 1 |
| Y | 3116 hCoV-19/Wales/PHWC-495831/2020\|EPI_ISL_726403\|2020 | ···················· | ···················· | ···················· | ···················· | NNNNNNNNNNNNNNNNNNNN | NNNNNNNNNNNNNNNNNNNN | 1 |
| Y | 3118 hCoV-19/Wales/PHWC-49585F/2020\|EPI_ISL_726405\|2020 | ···················· | ···················· | ···················· | ···················· | NNNNNNNNNNNNNNNNNNNN | NNNNNNNNNNNNNNNNNNNN | 1 |
| Y | 3119 hCoV-19/Wales/PHWC-49586E/2020\|EPI_ISL_726406\|2020 | ···················· | ···················· | ···················· | ···················· | NNNNNNNNNNNNNNNNNNNN | NNNNNNNNNNNNNNNNNNNN | 1 |
| Y | 3120 hCoV-19/Wales/PHWC-4958C8/2020\|EPI_ISL_726410\|2020 | ···················· | ···················· | ···················· | ···················· | NNNNNNNNNNNNNNNNNNNN | ···················· | 1 |
| Y | 3124 hCoV-19/Wales/PHWC-49592F/2020\|EPI_ISL_726415\|2020 | ···················· | ···················· | ···················· | ···················· | NNNNNNNNNNNNNNNNNNNN | NNNNNNNNNNNNNNNNNNNN | 1 |
| Y | 3128 hCoV-19/Wales/PHWC-49597A/2020\|EPI_ISL_726419\|2020 | ···················· | ···················· | ···················· | ···················· | NNNNNNNNNNNNNNNNNNNN | NNN················· | 1 |
| Y | 3129 hCoV-19/Wales/PHWC-495989/2020\|EPI_ISL_726420\|2020 | ···················· | ···················· | ···················· | ···················· | NNNNNNNNNNNNNNNNNNNN | NNNNNNNNNNNNNNNNNNNN | 1 |
| Y | 3132 hCoV-19/Wales/PHWC-4959C5/2020\|EPI_ISL_726423\|2020 | ···················· | ···················· | ···················· | ···················· | NNNNNNNNNNNNNNNNNNNN | ···················· | 1 |
| Y | 3133 hCoV-19/Wales/PHWC-4959D4/2020\|EPI_ISL_726424\|2020 | ···················· | ···················· | ···················· | ···················· | ···················· | NNNNNNNNNNN········· | 1 |

| | | | | | | | | | |
|---|---|---|---|---|---|---|---|---|---|
| Y | 3135 | hCoV-19/Wales/PHWC-495A0E/2020\|EPI_ISL_726426\|2020 | ···················· | ···················· | ···················· | ···················· | NNNNNNNNNNNNNNNNNNNN | NNNNNNNNNNNNNNNNNNNN | 1 |
| Y | 3136 | hCoV-19/Wales/PHWC-495A2C/2020\|EPI_ISL_726428\|2020 | ···················· | ·····A·············· | ···················· | ···················· | NNNNNNNNNNNNNNNNNNNN | NNNNNNNNNNNNNNNNNNNN | 1 |
| Y | 3139 | hCoV-19/Wales/PHWC-495AB3/2020\|EPI_ISL_726435\|2020 | ···················· | ···················· | ···················· | ···················· | ···················· | NNNNNNNNNNNNNNNNNNNN | 1 |
| Y | 3143 | hCoV-19/Wales/PHWC-495AFF/2020\|EPI_ISL_726439\|2020 | ···················· | ···················· | ···················· | ···················· | NNNN··NNNNNNNNNNNNNN | NNNNNNNNNNNNNNNNNNNN | 1 |
| Y | 3146 | hCoV-19/Wales/PHWC-495B29/2020\|EPI_ISL_726442\|2020 | ···················· | ···················· | ···················· | ···················· | NNNNNNNNNNNNNNNNNNNN | NNNNNNNNNNNNNNNNNNNN | 1 |
| Y | 3149 | hCoV-19/Wales/PHWC-495B56/2020\|EPI_ISL_726445\|2020 | ···················· | ···················· | ···················· | ···················· | NNNNNNNNNNNNNNNNNNNN | ···················· | 1 |
| Y | 3150 | hCoV-19/Wales/PHWC-495B65/2020\|EPI_ISL_726446\|2020 | ···················· | ···················· | ···················· | ···················· | ···················· | NNNNNNNNNNN········· | 1 |
| Y | 3155 | hCoV-19/Wales/PHWC-495BB0/2020\|EPI_ISL_726451\|2020 | ···················· | ···················· | ···················· | ···················· | NNNNNNNNNNNNNNNNNNNN | ····NN···········NNN | 1 |
| Y | 3156 | hCoV-19/Wales/PHWC-495BCF/2020\|EPI_ISL_726452\|2020 | ···················· | ···················· | ···················· | ···················· | NNNN················ | ···················· | 1 |
| Y | 3159 | hCoV-19/Wales/PHWC-495BFC/2020\|EPI_ISL_726455\|2020 | ···················· | ···················· | ···················· | ···················· | NNNNNNNNNNNNNNNNNNNN | NNNNNNNNNNNNNNNNNNNN | 1 |
| Y | 3160 | hCoV-19/Wales/PHWC-495C08/2020\|EPI_ISL_726456\|2020 | ···················· | ···················· | ···················· | ···················· | NNNNNNNNNNNNNNNNNNNN | NNNNNNNNNNNNNNNNNNNN | 1 |
| Y | 3162 | hCoV-19/Wales/PHWC-495C35/2020\|EPI_ISL_726459\|2020 | ···················· | ···················· | ···················· | ···················· | NNNNNNNNNNNNNNNNNNNN | NNNNNNNNNNN···NNNNNN | 1 |
| Y | 3171 | hCoV-19/Wales/PHWC-495CCC/2020\|EPI_ISL_726468\|2020 | ···················· | ···················· | ···················· | ···················· | NNNNNNNNNNNNNNNNNNNN | ···················· | 1 |
| Y | 3220 | hCoV-19/Portugal/PT2044/2020\|EPI_ISL_732259\|2020-1 | ···················· | ···················· | ···················· | ···················· | NNNNNNNNNNNNNNNNNNNN | NNNNNNNNNNNNNNNNNNNN | 1 |
| Y | 3348 | hCoV-19/Wales/PHWC-495E20/2020\|EPI_ISL_726490\|2020 | ···················· | ···················· | ···················· | ···················· | ······NNNNNNNNNNNNNN | NNNNNNNNNNNNNNNNNNNN | 1 |
| Y | 3351 | hCoV-19/Wales/PHWC-495E5D/2020\|EPI_ISL_726493\|2020 | ···················· | ···················· | ···················· | ···················· | NNNNNNNNNNNNNNNNNNNN | ···················· | 1 |
| Y | 3353 | hCoV-19/Wales/PHWC-495E7B/2020\|EPI_ISL_726495\|2020 | ···················· | ···················· | ···················· | ···················· | NNNNNNNNNNNNNNNNNNNN | NNNNNNNNNNNNNNNNNNNN | 1 |
| Y | 3357 | hCoV-19/Wales/PHWC-495EB7/2020\|EPI_ISL_726499\|2020 | ···················· | ···················· | ···················· | ···················· | NNNNNNNNNNNNNNNNNNNN | NNNNNNNNNNNNNNNNNNNN | 1 |
| Y | 3532 | hCoV-19/USA/MI-MDHHS-SC22602/2020\|EPI_ISL_737175\|2 | ···················· | ···················· | ···················· | ···················· | NNNNNNNNNNNNNNNNNNNN | ···················· | 1 |
| Y | 3620 | hCoV-19/Wales/PHWC-495ED5/2020\|EPI_ISL_726501\|2020 | ···················· | ···················· | ···················· | ···················· | NNNNNNNNNNNNNNNNNNNN | NNNNNNNNNNNNNNNNNNNN | 1 |
| Y | 3621 | hCoV-19/Wales/PHWC-495EF3/2020\|EPI_ISL_726503\|2020 | ···················· | ···················· | ···················· | ···················· | NNNNNNNNNNNNNNNNNNNN | NNNNNNNNNNNNNNNNNNNN | 1 |
| Y | 3623 | hCoV-19/Wales/PHWC-495F1E/2020\|EPI_ISL_726505\|2020 | ···················· | ···················· | ···················· | ···················· | NNNNNNNNNNNNNNNNNNNN | ···················· | 1 |
| Y | 3625 | hCoV-19/Wales/PHWC-495F4B/2020\|EPI_ISL_726508\|2020 | ···················· | ···················· | ···················· | ···················· | NNNNNNNNNNNNNNNNNNNN | NNNNNNNNNNNNNNNNNNNN | 1 |
| Y | 3627 | hCoV-19/Wales/PHWC-495F69/2020\|EPI_ISL_726510\|2020 | ···················· | ···················· | ···················· | ···················· | NNNNNNNNNNNNNNNNNNNN | NNNNNNNNNNNNNNN····· | 1 |
| Y | 3628 | hCoV-19/Wales/PHWC-495F87/2020\|EPI_ISL_726512\|2020 | ···················· | ···················· | ···················· | ···················· | NNNNNNNNNNNNNNNNNNNN | NNNNNNNNNNNNNNNNNNNN | 1 |
| Y | 3631 | hCoV-19/Wales/PHWC-495FD2/2020\|EPI_ISL_726517\|2020 | ···················· | ···················· | ···················· | ···················· | NNNNNNNNNNNNNNNNNNNN | NNNNNNNNNNNNNNNNNNNN | 1 |
| Y | 3635 | hCoV-19/Wales/PHWC-496065/2020\|EPI_ISL_726523\|2020 | ···················· | ···················· | ···················· | ···················· | NNNNNNNNNNNNNNNNNNNN | NNNNNNNNNNNNNNN····· | 1 |
| Y | 3637 | hCoV-19/Wales/PHWC-496083/2020\|EPI_ISL_726525\|2020 | ···················· | ···················· | ···················· | ···················· | NNNNNNNNNNNNNNNNNNNN | NNNNNNNNNNNNNNNNNNNN | 1 |
| Y | 3638 | hCoV-19/Wales/PHWC-496092/2020\|EPI_ISL_726526\|2020 | ···················· | ···················· | ···················· | ···················· | ···················· | NNNNNNNNNNNNNNN····· | 1 |
| Y | 3640 | hCoV-19/Wales/PHWC-4960B0/2020\|EPI_ISL_726528\|2020 | ···················· | ···················· | ···················· | ···················· | NNNNNNNNNNNNNNNNNNNN | NNNNNNNNNNNNNNNNNNNN | 1 |
| Y | 3642 | hCoV-19/Wales/PHWC-4960DE/2020\|EPI_ISL_726530\|2020 | ···················· | ···················· | ···················· | ···················· | ···················· | NNNNNNNNNNNNNNNNNNNN | 1 |
| Y | 3644 | hCoV-19/Wales/PHWC-4960FC/2020\|EPI_ISL_726532\|2020 | ···················· | ···················· | ···················· | ···················· | NNNNNNNNNNNNNNNNNNNN | ···················· | 1 |
| Y | 3653 | hCoV-19/Wales/PHWC-49619F/2020\|EPI_ISL_726542\|2020 | ···················· | ···················· | ···················· | ···················· | NNNNNNNNNNNNNNNNNNNN | NNNNNNNNNNNNNNNNNNNN | 1 |
| Y | 3654 | hCoV-19/Wales/PHWC-4961CC/2020\|EPI_ISL_726543\|2020 | ···················· | ···················· | ···················· | ···················· | NNNN················ | ···················· | 1 |
| Y | 3662 | hCoV-19/Wales/PHWC-496250/2020\|EPI_ISL_726552\|2020 | ···················· | ···················· | ···················· | ···················· | NNNNN··············· | ···················· | 1 |
| Y | 3666 | hCoV-19/Wales/PHWC-4962AB/2020\|EPI_ISL_726556\|2020 | ···················· | ···················· | ···················· | ···················· | NNNNNNNNNNNNNNNNNNNN | NNNNNNNNNNNNNNNNNNNN | 1 |
| Y | 3670 | hCoV-19/Wales/PHWC-496302/2020\|EPI_ISL_726562\|2020 | ···················· | ···················· | ···················· | ···················· | NNNN··············NN | ···················· | 1 |
| Y | 3671 | hCoV-19/Wales/PHWC-496311/2020\|EPI_ISL_726563\|2020 | ···················· | ···················· | ···················· | ···················· | NNNNNNNNNNNNNNNNNNNN | NNNNNNNNNNNNNNNNNNNN | 1 |
| Y | 3672 | hCoV-19/Wales/PHWC-496320/2020\|EPI_ISL_726564\|2020 | ···················· | ···················· | ···················· | ···················· | NNNNNNNNNNNNNNNNN··· | ···················· | 1 |
| Y | 3687 | hCoV-19/Wales/PHWC-4964D2/2020\|EPI_ISL_726590\|2020 | ···················· | ···················· | ···················· | ···················· | NNNNNNNNNNNNNNNNNNNN | NNNNNNNNNNNNNNNNNNNN | 1 |
| Y | 3688 | hCoV-19/Wales/PHWC-4964E1/2020\|EPI_ISL_726591\|2020 | ···················· | ···················· | ···················· | ···················· | NNNNNNNNNNNNNNNNNNNN | NNNNNNNNNNNNNNNNNNNN | 1 |
| Y | 3691 | hCoV-19/Wales/PHWC-49652A/2020\|EPI_ISL_726595\|2020 | ···················· | ···················· | ···················· | ···················· | NNNNNNNNNNNNNNNNNNNN | NNNNNNNNNNNNNNN····· | 1 |
| Y | 3692 | hCoV-19/Wales/PHWC-496539/2020\|EPI_ISL_726596\|2020 | ···················· | ···················· | ···················· | ···················· | NNNNNNNNNNNNNNNNNNNN | ···················· | 1 |
| Y | 3694 | hCoV-19/Wales/PHWC-496557/2020\|EPI_ISL_726598\|2020 | ···················· | ···················· | ···················· | ···················· | NNNNNNNNNNNNNNNNNNNN | ···················· | 1 |
| Y | 3695 | hCoV-19/Wales/PHWC-496566/2020\|EPI_ISL_726599\|2020 | ···················· | ···················· | ···················· | ···················· | NNNNNNNNNNNNNNNNNNNN | NNNNNNNNNNNNNNNNNNNN | 1 |
| Y | 3808 | hCoV-19/Finland/13ES23H9/2020\|EPI_ISL_737210\|2020- | ···················· | ···················· | ···················· | ···················· | ······NNNNNNNNNNNNNN | NNNNNNNNNNNNNNNNNNNN | 1 |
| Y | 3862 | hCoV-19/England/NORW-F6E6D/2020\|EPI_ISL_725306\|202 | ···················· | ···················· | ···················· | ···················· | NNNNNNNNNNNNNNNNNNNN | NNNNNNNNNNNNNNNNNNNN | 1 |
| Y | 3873 | hCoV-19/Wales/PHWC-496469/2020\|EPI_ISL_726583\|2020 | ···················· | ···················· | ···················· | ···················· | NNNNNNNNNNNNNNNNNNNN | NNNNNNNNNNNNNNNNNNNN | 1 |
| Y | 3878 | hCoV-19/Wales/PHWC-4964B4/2020\|EPI_ISL_726588\|2020 | ···················· | ···················· | ···················· | ···················· | NNNNNNNNNNNNNNNNNNNN | ···················· | 1 |
| Y | 3879 | hCoV-19/Wales/PHWC-4964C3/2020\|EPI_ISL_726589\|2020 | ···················· | ···················· | ···················· | ···················· | NNNNNNNNNNNNNNNNNNNN | NNNNNNNNNNNNNNNNNNNN | 1 |
| Y | 3881 | hCoV-19/Wales/PHWC-496584/2020\|EPI_ISL_726601\|2020 | ···················· | ···················· | ···················· | ···················· | NNNNNNNNNNNNNNNNNNNN | NNNNNNNNNNNNNNNNNNNN | 1 |
| Y | 3884 | hCoV-19/Wales/PHWC-4965B1/2020\|EPI_ISL_726604\|2020 | ···················· | ···················· | ···················· | ···················· | NNNNNNNNNNNNNN······ | NNNNNNNN············ | 1 |
| Y | 3885 | hCoV-19/Wales/PHWC-4965C0/2020\|EPI_ISL_726605\|2020 | ···················· | ···················· | ···················· | ···················· | NNNNNNNNNNNNNNNNNNNN | NNNNNNNNNNNNNNNNNNNN | 1 |
| Y | 3891 | hCoV-19/Wales/PHWC-496742/2020\|EPI_ISL_726629\|2020 | ···················· | ···················· | ···················· | ···················· | NNNNNNNNNNNNNNNNNNNN | NNNNNNNNNNNNNNNNNNNN | 1 |
| Y | 3895 | hCoV-19/Wales/PHWC-4967E8/2020\|EPI_ISL_726638\|2020 | ···················· | ···················· | ···················· | ···················· | NNN····NNNNNNNNNNNNN | ···················· | 1 |
| Y | 3897 | hCoV-19/Wales/PHWC-496812/2020\|EPI_ISL_726641\|2020 | ···················· | ···················· | ···················· | ···················· | NNNNNNNNNNNNNNNNNNNN | NNNNNNNNNNNNNNNNNNNN | 1 |
| Y | 3900 | hCoV-19/Wales/PHWC-49685E/2020\|EPI_ISL_726645\|2020 | ···················· | ···················· | ···················· | ···················· | NNNNNNNNNNNNNNNNNNNN | NNNNNNNNNNNNNNNNNNNN | 1 |
| Y | 3901 | hCoV-19/Wales/PHWC-49686D/2020\|EPI_ISL_726646\|2020 | ···················· | ···················· | ···················· | ···················· | NNNNNNNNNNNNNNNNNNNN | NNNNNNNNNNNNNNNNNNNN | 1 |
| Y | 3903 | hCoV-19/Wales/PHWC-49688B/2020\|EPI_ISL_726648\|2020 | ···················· | ···················· | ···················· | ···················· | NNNNNNNNNNNNNNNNNNNN | NNNNNNNNNNNNNNNNNNNN | 1 |
| Y | 4019 | hCoV-19/Ireland/D-NVRL-79IRL07665/2020\|EPI_ISL_732 | ···················· | ···················· | ···················· | ···················· | NNNNNNNNNNNNNNNNNNNN | NNNNNNNNNNNNNNNNNNNN | 1 |
| Y | 4105 | hCoV-19/England/NORW-F75B2/2020\|EPI_ISL_725335\|202 | ···················· | ···················· | ···················· | ···················· | NNNNNNNNNNNNNNNNNNNN | NNNNNNNNNNNNNNNNNNNN | 1 |
| Y | 4109 | hCoV-19/England/NORW-F76EC/2020\|EPI_ISL_725340\|202 | ···················· | ···················· | ···················· | ···················· | NNNNNNNNNNNNNNNNNNNN | ···················· | 1 |
| Y | 4139 | hCoV-19/Wales/PHWC-496627/2020\|EPI_ISL_726611\|2020 | ···················· | ···················· | ···················· | ···················· | NNNNNNNNNNNNNNNNNNNN | NNNNNNNNNNNNNNNNNNNN | 1 |
| Y | 4143 | hCoV-19/Wales/PHWC-4966CD/2020\|EPI_ISL_726621\|2020 | ···················· | ···················· | ···················· | ···················· | NNNNNNNNNNNNNNNNNNNN | ···················· | 1 |
| Y | 4152 | hCoV-19/Wales/PHWC-4968B8/2020\|EPI_ISL_726651\|2020 | ···················· | ···················· | ···················· | ···················· | NNNNNNNNNNNNNNNNNNNN | NNNNNNNNNNNNNNNNNNNN | 1 |
| Y | 4155 | hCoV-19/Wales/PHWC-4968F4/2020\|EPI_ISL_726655\|2020 | ···················· | ···················· | ···················· | ···················· | NNNNNNNNNNNNNNNNNNNN | NNNNN···········NNNN | 1 |
| Y | 4157 | hCoV-19/Wales/PHWC-49692E/2020\|EPI_ISL_726658\|2020 | ···················· | ·····A·············· | ···················· | ···················· | NNNNNNNNNNNNNNNNNNNN | NNNNNNNNNNNNNNNNNNNN | 1 |
| Y | 4158 | hCoV-19/Wales/PHWC-49694C/2020\|EPI_ISL_726659\|2020 | ···················· | ···················· | ···················· | ···················· | NNNNNNNNNNNNNNNNNNNN | NNNNNNNNNNNNNNNNNNNN | 1 |
| Y | 4159 | hCoV-19/Wales/PHWC-49695B/2020\|EPI_ISL_726660\|2020 | ···················· | ···················· | ···················· | ···················· | NNNNNNNNNNNNNNNNNNNN | NNNNNNNNNNNNNNNNNNNN | 1 |
| Y | 4160 | hCoV-19/Wales/PHWC-496988/2020\|EPI_ISL_726663\|2020 | ···················· | ···················· | ···················· | ···················· | NNNNNNNNNNNNNNNNNNNN | NNNNNNNNNNNNNNNNNNNN | 1 |
| Y | 4162 | hCoV-19/Wales/PHWC-4969C4/2020\|EPI_ISL_726667\|2020 | ···················· | ···················· | ···················· | ···················· | NNNNNNNNNNNNNNNNNNNN | ·····NNNNNNNNNN····· | 1 |
| Y | 4163 | hCoV-19/Wales/PHWC-4969D3/2020\|EPI_ISL_726668\|2020 | ···················· | ···················· | ···················· | ···················· | ······NNNNNN········ | ···················· | 1 |
| Y | 4164 | hCoV-19/Wales/PHWC-4969E2/2020\|EPI_ISL_726669\|2020 | ···················· | ···················· | ···················· | ···················· | NNNNNNNNNNNNNNNNNNNN | NNNNNNNNNNNNNNNNNNNN | 1 |
| Y | 4165 | hCoV-19/Wales/PHWC-4969F1/2020\|EPI_ISL_726670\|2020 | ···················· | ···················· | ···················· | ···················· | NNNNNNNNNNNNNNNNNNNN | NNNNNNNNNNNNNNNNNNNN | 1 |
| Y | 4166 | hCoV-19/Wales/PHWC-496A0D/2020\|EPI_ISL_726671\|2020 | ···················· | ···················· | ···················· | ···················· | NNNNNNNNNNNNNNNNNNNN | NNNNNNNNNNNNNNNNNNNN | 1 |
| Y | 4168 | hCoV-19/Wales/PHWC-496A2B/2020\|EPI_ISL_726673\|2020 | ···················· | ···················· | ···················· | ···················· | NNNNNNNNNNNNNNNNNNNN | NNNNNNNNNNNNNNNNNNNN | 1 |
| Y | 4170 | hCoV-19/Wales/PHWC-496A49/2020\|EPI_ISL_726675\|2020 | ···················· | ···················· | ···················· | ···················· | NNNNNNNNNNNNNNNNNNNN | NNNNNNNNNNNNNNNNNNNN | 1 |
| Y | 4173 | hCoV-19/Wales/PHWC-496A94/2020\|EPI_ISL_726680\|2020 | ···················· | ···················· | ···················· | ···················· | NNNNNNNNNNNNNNNNNNNN | NNNNNNNNNNNNNNNNNNNN | 1 |
| Y | 4174 | hCoV-19/Wales/PHWC-496AA3/2020\|EPI_ISL_726681\|2020 | ···················· | ···················· | ···················· | ···················· | NNNNNNNNNNNNNNNNNNNN | NNNNNNNNNNNNNNNNNNNN | 1 |
| Y | 4175 | hCoV-19/Wales/PHWC-496AC1/2020\|EPI_ISL_726683\|2020 | ···················· | ···················· | ···················· | ···················· | NNNNNNNNNNNNNNNNNNNN | NNNNNNNNNNNNNNNNNNNN | 1 |

| | | | | | | | | | | | |
|---|---|---|---|---|---|---|---|---|---|---|---|
| Y | 4177 | hCoV-19/Wales/PHWC-496AEF/2020 | EPI_ISL_726685 | 2020 | ···················· | ···················· | ···················· | ···················· | NNNNNNNNNNNNNNNNNNNN | NNNNNNNNNNNNNNNNNNNN | 1 |
| Y | 4178 | hCoV-19/Wales/PHWC-496AFE/2020 | EPI_ISL_726686 | 2020 | ···················· | ···················· | ···················· | ···················· | NNNNNNNNNNNNNNNNNNNN | NNNNNNNNNNNNNNNNNNNN | 1 |
| Y | 4182 | hCoV-19/Wales/PHWC-496B46/2020 | EPI_ISL_726691 | 2020 | ···················· | ···················· | ···················· | ···················· | NNNNNNNNNNNNNNNNNNNN | NNNNNNNNNNNNNNNNNNNN | 1 |
| Y | 4186 | hCoV-19/Wales/PHWC-496B82/2020 | EPI_ISL_726695 | 2020 | ···················· | ···················· | ···················· | ···················· | NNNNNNNNNNNNNNNNNNNN | NNNNNNNNNNNNNNNNNNNN | 1 |
| Y | 4188 | hCoV-19/Wales/PHWC-496BCE/2020 | EPI_ISL_726698 | 2020 | ···················· | ···················· | ···················· | ···················· | NNNNNNNNNNNNNNNNNNNN | NNNNNNNNNNNNNNNNNNNN | 1 |
| Y | 4296 | hCoV-19/USA/MI-MDHHS-SC22687/2020 | EPI_ISL_737301 | 2 | ···················· | ···················· | ···················· | ···················· | NNNNNN··NNNN··N····· | ···················· | 1 |
| Y | 4360 | hCoV-19/England/NORW-F992D/2020 | EPI_ISL_725410 | 202 | ···················· | ···················· | ···················· | ···················· | NNNNNNNNNNNNNNNNNNNN | NNNNN············NNN | 1 |
| Y | 4361 | hCoV-19/England/NORW-F994B/2020 | EPI_ISL_725412 | 202 | ···················· | ···················· | ···················· | ···················· | ···················· | NNNNNNNNNNNNNNNNNNNN | 1 |
| Y | 4381 | hCoV-19/Wales/PHWC-496BEC/2020 | EPI_ISL_726700 | 2020 | ···················· | ···················· | ···················· | ···················· | NNNNNNNNNNNNNNNNNNNN | NNNNNNNNNNNNNNNNNNNN | 1 |
| Y | 4382 | hCoV-19/Wales/PHWC-496C07/2020 | EPI_ISL_726702 | 2020 | ···················· | ···················· | ···················· | ···················· | NNNNNNNNNNNNNNNNNNNN | NNNNNNNNNNNNNNNNNNNN | 1 |
| Y | 4384 | hCoV-19/Wales/PHWC-496C25/2020 | EPI_ISL_726704 | 2020 | ···················· | ···················· | ···················· | ···················· | NNNNNNNNNNNNNNNNNNNN | NNNNNNNNNNNNNNNNNNNN | 1 |
| Y | 4387 | hCoV-19/Wales/PHWC-496C52/2020 | EPI_ISL_726707 | 2020 | ···················· | ···················· | ···················· | ···················· | NNNNNNNNNNNNNNNNNNNN | NNNNNNNNNNNNNNNNNNNN | 1 |
| Y | 4390 | hCoV-19/Wales/PHWC-496CAD/2020 | EPI_ISL_726711 | 2020 | ···················· | ·····A·············· | ···················· | ···················· | NNNNNNNNNNNNNNNNNNNN | ···················· | 1 |
| Y | 4392 | hCoV-19/Wales/PHWC-496CCB/2020 | EPI_ISL_726713 | 2020 | ···················· | ···················· | ···················· | ···················· | NNNNNNNNNNNNNNNNNNNN | ···················· | 1 |
| Y | 4397 | hCoV-19/Wales/PHWC-496D22/2020 | EPI_ISL_726719 | 2020 | ···················· | ···················· | ···················· | ···················· | NNNNNNNNNNNNNNNNNNNN | NNNNNNNNNNNNNNNNNNNN | 1 |
| Y | 4405 | hCoV-19/Wales/PHWC-496DAA/2020 | EPI_ISL_726727 | 2020 | ···················· | ···················· | ···················· | ···················· | NNNNNNNNNNNNNNNNNNNN | NNNNNNNNNNNNNNNNNNNN | 1 |
| Y | 4409 | hCoV-19/Wales/PHWC-496DF5/2020 | EPI_ISL_726732 | 2020 | ···················· | ···················· | ···················· | ···················· | NNNNNNNNNNNNNNNNNNNN | ···················· | 1 |
| Y | 4415 | hCoV-19/Wales/PHWC-496E6B/2020 | EPI_ISL_726738 | 2020 | ···················· | ···················· | ···················· | ···················· | NNNNNNNNNNNNNNNNNNNN | NNNNNNNNNNNNNNNNNNNN | 1 |
| Y | 4416 | hCoV-19/Wales/PHWC-496E7A/2020 | EPI_ISL_726739 | 2020 | ···················· | ···················· | ···················· | ···················· | ···················· | NNNNNNNNNNNNNNN····· | 1 |
| Y | 4420 | hCoV-19/Wales/PHWC-496EC5/2020 | EPI_ISL_726744 | 2020 | ···················· | ···················· | ···················· | ···················· | NNNNNNNNNNNNNNNNNNNN | NNNNNNNNNNNNNNNNNNNN | 1 |
| Y | 4421 | hCoV-19/Wales/PHWC-496ED4/2020 | EPI_ISL_726745 | 2020 | ···················· | ···················· | ···················· | ···················· | NNNNNNNNNNNNNNNNNNNN | ·········NNNNNN····N | 1 |
| Y | 4422 | hCoV-19/Wales/PHWC-496EE3/2020 | EPI_ISL_726746 | 2020 | ···················· | ···················· | ···················· | ···················· | NNNNNNNNNNNNNNNNNNNN | ···NNNNNNNN······NNN | 1 |
| Y | 4425 | hCoV-19/Wales/PHWC-496F2C/2020 | EPI_ISL_726750 | 2020 | ···················· | ···················· | ···················· | ···················· | ···················· | NNNNNNNNNNNNNNN····· | 1 |
| Y | 4430 | hCoV-19/Wales/PHWC-496F86/2020 | EPI_ISL_726756 | 2020 | ···················· | ···················· | ···················· | ···················· | ···················· | NNNNNNNNNNNNNNNNNNNN | 1 |
| Y | 4435 | hCoV-19/Wales/PHWC-496FE0/2020 | EPI_ISL_726762 | 2020 | ···················· | ···················· | ···················· | ···················· | NNNNNNNNNNNNNNNNNNNN | NNNNNNNNNNNNNNNNNNNN | 1 |
| Y | 4436 | hCoV-19/Wales/PHWC-496FFF/2020 | EPI_ISL_726763 | 2020 | ···················· | ···················· | ···················· | ···················· | NNNNNNNNNNNNNNNNNNNN | ···················· | 1 |
| Y | 4437 | hCoV-19/Wales/PHWC-49700A/2020 | EPI_ISL_726764 | 2020 | ···················· | ···················· | ···················· | ···················· | ···················· | NNNNNNNNNNNNNNN····· | 1 |
| Y | 4439 | hCoV-19/Wales/PHWC-497028/2020 | EPI_ISL_726766 | 2020 | ···················· | ···················· | ···················· | ···················· | ···················· | NNNNNNNNNNNNNNNNNNNN | 1 |
| Y | 4440 | hCoV-19/Wales/PHWC-497037/2020 | EPI_ISL_726767 | 2020 | ···················· | ···················· | ···················· | ···················· | ···················· | ···········NNNN····· | 1 |
| Y | 4450 | hCoV-19/Wales/PHWC-497213/2020 | EPI_ISL_726797 | 2020 | ···················· | ···················· | ···················· | ···················· | NNNNNNNNNNNNNNNNNNNN | ···················· | 1 |
| Y | 4451 | hCoV-19/Wales/PHWC-497222/2020 | EPI_ISL_726798 | 2020 | ···················· | ···················· | ···················· | ···················· | NNNNNNNNNNNNNNNNNNNN | NNNNNNNNNNNNNNNNNNNN | 1 |
| Y | 4452 | hCoV-19/Wales/PHWC-497231/2020 | EPI_ISL_726799 | 2020 | ···················· | ···················· | ···················· | ···················· | ···················· | NNNNNNNNNNNNNNN····· | 1 |
| Y | 4609 | hCoV-19/England/NORW-F9CE8/2020 | EPI_ISL_725440 | 202 | ···················· | ···················· | ···················· | ···················· | NNNNN··············· | NNNNNNNNNNNNNNNNNNNN | 1 |
| Y | 4628 | hCoV-19/England/NORW-FA35B/2020 | EPI_ISL_725497 | 202 | ···················· | ···················· | ···················· | ···················· | NNNNNNNNNNNNNNNNNNNN | NNNNNNNNNNNNNNNNNNNN | 1 |
| Y | 4632 | hCoV-19/Wales/PHWC-497091/2020 | EPI_ISL_726773 | 2020 | ···················· | ···················· | ···················· | ···················· | NNNNNNNNNNNNNNNNNNNN | NNNNNNNNNNNNNNNNNNNN | 1 |
| Y | 4633 | hCoV-19/Wales/PHWC-4970A0/2020 | EPI_ISL_726774 | 2020 | ···················· | ···················· | ···················· | ···················· | NNNNNNNNNNNNNNNNNNNN | NNNNN··············· | 1 |
| Y | 4634 | hCoV-19/Wales/PHWC-4970BF/2020 | EPI_ISL_726775 | 2020 | ···················· | ···················· | ···················· | ···················· | NNNNNNNNNNNNNNNNNNNN | NNNNNNNNNNNNNNNNNNNN | 1 |
| Y | 4635 | hCoV-19/Wales/PHWC-4970CE/2020 | EPI_ISL_726776 | 2020 | ···················· | ···················· | ···················· | ···················· | NNNNNNNNNNNNNNNNNNNN | NNNNNNNNNNNNNNNNNNNN | 1 |
| Y | 4636 | hCoV-19/Wales/PHWC-4970DD/2020 | EPI_ISL_726777 | 2020 | ···················· | ···················· | ···················· | ···················· | NNNNNNNNNNNNNNNNNNNN | NNNNNNNNNNNNNNNNNNNN | 1 |
| Y | 4637 | hCoV-19/Wales/PHWC-4970EC/2020 | EPI_ISL_726778 | 2020 | ···················· | ···················· | ···················· | ···················· | NNNNNNNNNNNNNNNNNNNN | NNNNNNNNNNNNNNNNNNNN | 1 |
| Y | 4638 | hCoV-19/Wales/PHWC-4970FB/2020 | EPI_ISL_726779 | 2020 | ···················· | ···················· | ···················· | ···················· | NNNNNNNNNNNNNNNNNNNN | NNNNNNNNNNNNNNNNNNNN | 1 |
| Y | 4642 | hCoV-19/Wales/PHWC-497134/2020 | EPI_ISL_726783 | 2020 | ···················· | ···················· | ···················· | ···················· | NNNNNNNNNNNNNNNNNNNN | NNNNNNNNNNN·····NNNN | 1 |
| Y | 4652 | hCoV-19/Wales/PHWC-498EE1/2020 | EPI_ISL_727227 | 2020 | ···················· | ···················· | ···················· | ···················· | NNNNNNNNNNNNNNNNNNNN | NNNNNNNNNNNNNNNNNNNN | 1 |
| Y | 4653 | hCoV-19/Wales/PHWC-498EF0/2020 | EPI_ISL_727228 | 2020 | ···················· | ···················· | ···················· | ···················· | NNNN················ | ···················· | 1 |
| Y | 4654 | hCoV-19/Wales/PHWC-498F0C/2020 | EPI_ISL_727229 | 2020 | ···················· | ···················· | ···················· | ···················· | NNNNNNNNNNNNNNNNNNNN | NNNNNNNNNNNNNNNNNNNN | 1 |
| Y | 4656 | hCoV-19/Wales/PHWC-498F57/2020 | EPI_ISL_727233 | 2020 | ···················· | ·····A·············· | ···················· | ···················· | NNNNNNNNNNNNNNNNNNNN | NNNNNNNNNNNNNNNNNNNN | 1 |
| Y | 4657 | hCoV-19/Wales/PHWC-498F66/2020 | EPI_ISL_727234 | 2020 | ···················· | ···················· | ···················· | ···················· | NNNNNNNNNNNN········ | ···················· | 1 |
| Y | 4658 | hCoV-19/Wales/PHWC-498F75/2020 | EPI_ISL_727235 | 2020 | ···················· | ···················· | ···················· | ···················· | NNNNNNNNNNNNNNNNNNNN | NNNNNNNNNNNNNNNNNNNN | 1 |
| Y | 4659 | hCoV-19/Wales/PHWC-498F84/2020 | EPI_ISL_727236 | 2020 | ···················· | ···················· | ···················· | ···················· | NNNNNNNNNNNNNNNNNNNN | NNNNNNNNNNNNNNNNNNNN | 1 |
| Y | 4660 | hCoV-19/Wales/PHWC-498FA2/2020 | EPI_ISL_727238 | 2020 | ···················· | ···················· | ···················· | ···················· | NNNNNNNNNNNNNNNNNNNN | ···················· | 1 |
| Y | 4661 | hCoV-19/Wales/PHWC-498FB1/2020 | EPI_ISL_727239 | 2020 | ···················· | ···················· | ···················· | ···················· | NNNNNNNNNNNNNNNNNNNN | NNNNNNNNNNNNNNNNNNNN | 1 |
| Y | 4662 | hCoV-19/Wales/PHWC-498FC0/2020 | EPI_ISL_727240 | 2020 | ···················· | ···················· | ···················· | ···················· | NNNNNNNNNNNNNNNNNNNN | NNNNNNNNNNNNNNNNNNNN | 1 |
| Y | 4664 | hCoV-19/Wales/PHWC-499008/2020 | EPI_ISL_727244 | 2020 | ···················· | ···················· | ···················· | ···················· | NNNNNNNNNNNNNNNNNNNN | NNNNNNNNNNNNNNN····· | 1 |
| Y | 4665 | hCoV-19/Wales/PHWC-499017/2020 | EPI_ISL_727245 | 2020 | ···················· | ·····A·············· | ···················· | ···················· | NNNNNNNNNNNNNNNNNNNN | NNNNNNNNNNNNNNNNNNNN | 1 |
| Y | 4667 | hCoV-19/Wales/PHWC-499053/2020 | EPI_ISL_727249 | 2020 | ···················· | ···················· | ···················· | ···················· | ··NNNNNNNNNNNNNNNNN· | NNNNNNNNNNNNNNN····· | 1 |
| Y | 4668 | hCoV-19/Wales/PHWC-499062/2020 | EPI_ISL_727250 | 2020 | ···················· | ···················· | ···················· | ···················· | NNNNNNNNNNNNNNNNNNNN | ···················· | 1 |
| Y | 4669 | hCoV-19/Wales/PHWC-499071/2020 | EPI_ISL_727251 | 2020 | ···················· | ···················· | ···················· | ···················· | NNNNNNNNNNNNNNNNNNNN | ·········NNNNNNNNNNN | 1 |
| Y | 4670 | hCoV-19/Wales/PHWC-499080/2020 | EPI_ISL_727252 | 2020 | ···················· | ···················· | ···················· | ···················· | NNNNNNNNNNNNNNNNNNNN | NNNNNNNNNNNNNNNNNNNN | 1 |
| Y | 4672 | hCoV-19/Wales/PHWC-4990AE/2020 | EPI_ISL_727254 | 2020 | ···················· | ···················· | ···················· | ···················· | ···················· | NNNNNNNNNNNNNNN····· | 1 |
| Y | 4673 | hCoV-19/Wales/PHWC-4990DB/2020 | EPI_ISL_727255 | 2020 | ···················· | ···················· | ···················· | ···················· | NNNNNNNNNNNNNNNNNNNN | NNNNNNNNNNNNNNNNNNNN | 1 |
| Y | 4674 | hCoV-19/Wales/PHWC-4990EA/2020 | EPI_ISL_727256 | 2020 | ···················· | ···················· | ···················· | ···················· | ···················· | NNNNN··············· | 1 |
| Y | 4676 | hCoV-19/Wales/PHWC-499114/2020 | EPI_ISL_727258 | 2020 | ···················· | ···················· | ···················· | ···················· | NNNNNNNNNNNNNNNNNNNN | NNNNNNNNNNNNNNNNNNNN | 1 |
| Y | 4677 | hCoV-19/Wales/PHWC-499132/2020 | EPI_ISL_727260 | 2020 | ···················· | ·····A·············· | ···················· | ···················· | NNNNNNNNNNNNNNNNNNNN | NNNNNNNNNNNNNNNNNNNN | 1 |
| Y | 4678 | hCoV-19/Wales/PHWC-499141/2020 | EPI_ISL_727261 | 2020 | ···················· | ·····A·············· | ···················· | ···················· | NNNNNNNNNNNNNNNNNNNN | NNNNNNNNNNNNNNNNNNNN | 1 |
| Y | 4679 | hCoV-19/Wales/PHWC-499150/2020 | EPI_ISL_727262 | 2020 | ···················· | ·····A·············· | ···················· | ···················· | NNNNNNNNNNNNNNNNNNNN | NNNNNNNNNNNNNNNNNNNN | 1 |
| Y | 4680 | hCoV-19/Wales/PHWC-49918D/2020 | EPI_ISL_727265 | 2020 | ···················· | ···················· | ···················· | ···················· | NNNNNNNNNNNNNNNNNNNN | NNNNNNNNNNNNNNNNNNNN | 1 |
| Y | 4683 | hCoV-19/Wales/PHWC-4991C9/2020 | EPI_ISL_727269 | 2020 | ···················· | ·····A·············· | ···················· | ···················· | NNNNNNNNNNNNNNNNNNNN | NNNNNNNNNNN········· | 1 |
| Y | 4686 | hCoV-19/Wales/PHWC-499202/2020 | EPI_ISL_727272 | 2020 | ···················· | ···················· | ···················· | ···················· | NNNNNNNNNNNNNNNNNNNN | NNNNNNNNNNNNNNNNNNNN | 1 |
| Y | 4687 | hCoV-19/Wales/PHWC-499211/2020 | EPI_ISL_727273 | 2020 | ···················· | ···················· | ···················· | ···················· | NNNNNNNNNNNNNNNNNNNN | NNNNNNNNNNNNNNNNNNNN | 1 |
| Y | 4691 | hCoV-19/Wales/PHWC-49925D/2020 | EPI_ISL_727277 | 2020 | ···················· | ···················· | ···················· | ···················· | NNNNNNNNNNNNNNNNNNNN | NNNNN··············· | 1 |
| Y | 4692 | hCoV-19/Wales/PHWC-49926C/2020 | EPI_ISL_727278 | 2020 | ···················· | ···················· | ···················· | ···················· | NNNNNNNNNNNNNNNNNNNN | NNNNNNNNNNNNNNNNNNNN | 1 |
| Y | 4696 | hCoV-19/Wales/PHWC-4992B7/2020 | EPI_ISL_727283 | 2020 | ···················· | ···················· | ···················· | ···················· | NNNNNNNNNNNNNNNNNNNN | NNNNNNNNNNNNNNNNNNNN | 1 |
| Y | 4701 | hCoV-19/Wales/PHWC-49930F/2020 | EPI_ISL_727288 | 2020 | ···················· | ···················· | ···················· | ···················· | ···················· | NNNNN··············· | 1 |
| Y | 4703 | hCoV-19/Wales/PHWC-49933C/2020 | EPI_ISL_727291 | 2020 | ···················· | ···················· | ···················· | ···················· | NNNNNNNNNNNNNNNNNNNN | NNNNNNNNNNNNNNNNNNNN | 1 |
| Y | 4704 | hCoV-19/Wales/PHWC-49934B/2020 | EPI_ISL_727292 | 2020 | ···················· | ···················· | ···················· | ···················· | NNNNNNNNNNNNNNNNNNNN | NNNNNNNNNNNNNNNNNNNN | 1 |
| Y | 4705 | hCoV-19/Wales/PHWC-49935A/2020 | EPI_ISL_727293 | 2020 | ···················· | ···················· | ···················· | ···················· | NNNNNNNNNNNNNNNNNNNN | NNNNNNNNNNNNNNN····· | 1 |
| Y | 4708 | hCoV-19/Wales/PHWC-499396/2020 | EPI_ISL_727296 | 2020 | ···················· | ···················· | ···················· | ···················· | NNNN················ | NNNNNNNNNNNNNNNNNNNN | 1 |
| Y | 4709 | hCoV-19/Wales/PHWC-4993A5/2020 | EPI_ISL_727297 | 2020 | ···················· | ···················· | ···················· | ···················· | NNNNNNNNNNNNNNNNNNNN | NNNNNNNNNNNNNNNNNNNN | 1 |
| Y | 4711 | hCoV-19/Wales/PHWC-4993D2/2020 | EPI_ISL_727299 | 2020 | ···················· | ···················· | ···················· | ···················· | NNNNNNNNNNNNNNNNNNNN | NNNNNNNNNNNNNNNNNNNN | 1 |
| Y | 4865 | hCoV-19/Wales/PHWC-499545/2020 | EPI_ISL_727320 | 2020 | ···················· | ···················· | ···················· | ···················· | NNNNNNNNNNNNNNNNNNNN | NNNNNNNNNNNNNNNNNNNN | 1 |

| | | | | | | | | | |
|---|---|---|---|---|---|---|---|---|---|
| Y | 4867 | hCoV-19/Wales/PHWC-499581/2020\|EPI_ISL_727323\|2020 | ···················· | ···················· | ···················· | ···················· | ···················· | NNNNNNNNNNNNNNN····· | 1 |
| Y | 4872 | hCoV-19/Wales/PHWC-4995DC/2020\|EPI_ISL_727328\|2020 | ···················· | ···················· | ···················· | ···················· | NNNNNNNNNNNNNNNNNNNN | ·········NNNNNNNNNNN | 1 |
| Y | 5025 | hCoV-19/Russia/MOW-RII-MH4105S/2020\|EPI_ISL_733163 | ···················· | ···················· | ···················· | ···················· | ···················· | ················NNNN | 1 |
| Y | 5027 | hCoV-19/Russia/AST-RII-MH4131S/2020\|EPI_ISL_733168 | ···················· | ···················· | ···················· | ···················· | ···················· | NNNNNNNNNNNNNNNNNNNN | 1 |
| Y | 5029 | hCoV-19/Russia/AST-RII-MH4150S/2020\|EPI_ISL_733174 | ···················· | ···················· | ···················· | ···················· | NNNNNNNNNNNNNNNNNNNN | NNNNNNNNNNNNNNNNN··· | 1 |
| Y | 5039 | hCoV-19/Russia/AST-RII-MH4447S/2020\|EPI_ISL_733191 | ···················· | ···················· | ···················· | ···················· | NNNNNNNNNNNNNNNNNNNN | ···················· | 1 |
| Y | 5040 | hCoV-19/Russia/AST-RII-MH4449S/2020\|EPI_ISL_733192 | ···················· | ···················· | ···················· | ···················· | NNNNNNNNNNNNNNNNNNNN | ···················· | 1 |
| Y | 5041 | hCoV-19/Russia/AST-RII-MH4453S/2020\|EPI_ISL_733194 | ···················· | ···················· | ···················· | ···················· | ···················· | NNNNNNNNNNNNNNNNNNNN | 1 |
| Y | 5042 | hCoV-19/Russia/AST-RII-MH4455S/2020\|EPI_ISL_733196 | ···················· | ···················· | ···················· | ···················· | ···················· | NNNNNNNNNNNNNNNNNNNN | 1 |
| Y | 5045 | hCoV-19/Russia/AST-RII-MH4458S/2020\|EPI_ISL_733199 | ···················· | ···················· | ···················· | ···················· | NNNNNNNNNNNNNNNNNNNN | ···················· | 1 |
| Y | 5126 | hCoV-19/Wales/PHWC-493CCE/2020\|EPI_ISL_726010\|2020 | ···················· | ···················· | ···················· | ···················· | ···················· | NNNNN··············· | 1 |
| Y | 5139 | hCoV-19/Wales/PHWC-493DAD/2020\|EPI_ISL_726024\|2020 | ···················· | ···················· | ···················· | ···················· | NNNNNNNNNNNNNNNNNNNN | NNNNNNNNNNNNNNNNNNNN | 1 |
| Y | 5140 | hCoV-19/Wales/PHWC-493DCB/2020\|EPI_ISL_726026\|2020 | ···················· | ···················· | ···················· | ···················· | NNNNNNNNNNNNNNNNNNNN | NNNNNNNNNNNNNNNNNNNN | 1 |
| Y | 5143 | hCoV-19/Wales/PHWC-493E04/2020\|EPI_ISL_726030\|2020 | ···················· | ···················· | ···················· | ···················· | NNNNNNNNNNNNNNNNNNNN | NNNNNNNNNNN······NNN | 1 |
| Y | 5147 | hCoV-19/Wales/PHWC-493EC8/2020\|EPI_ISL_726036\|2020 | ···················· | ···················· | ···················· | ···················· | NNNNNNN············· | ···················· | 1 |
| Y | 5155 | hCoV-19/Wales/PHWC-493F6B/2020\|EPI_ISL_726045\|2020 | ···················· | ···················· | ···················· | ···················· | NNNNNNNNNNNN········ | ···················· | 1 |
| Y | 5161 | hCoV-19/Wales/PHWC-494191/2020\|EPI_ISL_726078\|2020 | ···················· | ···················· | ···················· | ···················· | NNNNNNN············· | NNNNNNNNNNNNNNN····· | 1 |
| Y | 5162 | hCoV-19/Wales/PHWC-4941A0/2020\|EPI_ISL_726079\|2020 | ···················· | ···················· | ···················· | ···················· | NNNN················ | NNNNNNNNNNN········· | 1 |
| Y | 5167 | hCoV-19/Wales/PHWC-499439/2020\|EPI_ISL_727304\|2020 | ···················· | ·····A·············· | ···················· | ···················· | NNNNNNNNNNNNNNNNNNNN | NNNNNNNNNNNNNNNNNNNN | 1 |
| Y | 5175 | hCoV-19/Wales/PHWC-4994C0/2020\|EPI_ISL_727313\|2020 | ···················· | ···················· | ···················· | ···················· | NNNNNNNNNNNNNNNNNNNN | NNNNNNNNNNNNNNNNNNNN | 1 |
| Y | 5177 | hCoV-19/Wales/PHWC-4994EE/2020\|EPI_ISL_727315\|2020 | ···················· | ···················· | ···················· | ···················· | ···················· | NNNNNNNNNNNNNNNNNNNN | 1 |
| Y | 5189 | hCoV-19/Wales/PHWC-4996BB/2020\|EPI_ISL_727341\|2020 | ···················· | ···················· | ···················· | ···················· | NNNNNNNNNNNNNNNNNNNN | NNNNNNNNNNNNNNNNNNNN | 1 |
| Y | 5191 | hCoV-19/Wales/PHWC-4996F7/2020\|EPI_ISL_727344\|2020 | ···················· | ···················· | ···················· | ···················· | ···················· | NNNNNNNNNNNNNNNNNNNN | 1 |
| Y | 5193 | hCoV-19/Wales/PHWC-499712/2020\|EPI_ISL_727346\|2020 | ···················· | ···················· | ···················· | ···················· | NNNNNNNNNNNNNNNNNNNN | NNNNNNNNNNNNNNNNNNNN | 1 |
| Y | 5200 | hCoV-19/Wales/PHWC-4997A9/2020\|EPI_ISL_727355\|2020 | ·T·················· | ···················· | ···················· | ···················· | NNNNNNNNNNNNNNNNNNNN | NNNNNNNNNNNNNNNNNNNN | 1 |
| Y | 5203 | hCoV-19/Wales/PHWC-4997D6/2020\|EPI_ISL_727358\|2020 | ···················· | ···················· | ···················· | ···················· | NNNNNNNNNNNNNNNNNNNN | NNNNNNNNNNNNNNNNNNNN | 1 |
| Y | 5207 | hCoV-19/Wales/PHWC-49981F/2020\|EPI_ISL_727362\|2020 | ···················· | ···················· | ···················· | ···················· | ···················· | NNNNNNNNNN·········· | 1 |
| Y | 5209 | hCoV-19/Wales/PHWC-49983D/2020\|EPI_ISL_727364\|2020 | ···················· | ···················· | ···················· | ···················· | NNNNNNNNNNNNNNNNNNNN | ···················· | 1 |
| Y | 5213 | hCoV-19/Wales/PHWC-499879/2020\|EPI_ISL_727368\|2020 | ···················· | ···················· | ···················· | ···················· | NNNNNNNNNNNNNNNNNNNN | ···················· | 1 |
| Y | 5215 | hCoV-19/Wales/PHWC-499897/2020\|EPI_ISL_727370\|2020 | ···················· | ·····A·············· | ···················· | ···················· | NNNNNNNNNNNNNNNNNNNN | NNNNNNNNNNNNNNNNNNNN | 1 |
| Y | 5216 | hCoV-19/Wales/PHWC-4998B5/2020\|EPI_ISL_727371\|2020 | ···················· | ···················· | ···················· | ···················· | NNNNNNNNNNNNNNNNNNNN | ···················· | 1 |
| Y | 5217 | hCoV-19/Wales/PHWC-4998C4/2020\|EPI_ISL_727372\|2020 | ···················· | ···················· | ···················· | ···················· | NNNNNNNNNNNNNNNNNNNN | NNNNNNNNNNNNNNNNNNNN | 1 |
| Y | 5220 | hCoV-19/Wales/PHWC-49992B/2020\|EPI_ISL_727378\|2020 | ···················· | ···················· | ···················· | ···················· | ···················· | NNNNNNNNNNNNNNN····· | 1 |
| Y | 5378 | hCoV-19/Wales/PHWC-493FE3/2020\|EPI_ISL_726052\|2020 | ···················· | ···················· | ···················· | ···················· | NNNNNNNNNNNNNNNNNNNN | NNNNNNNNNNNNNNNNNNNN | 1 |
| Y | 5379 | hCoV-19/Wales/PHWC-493FF2/2020\|EPI_ISL_726053\|2020 | ···················· | ···················· | ···················· | ···················· | NNNNNNNNNNNNNNNNNNNN | ··············NNNNNN | 1 |
| Y | 5382 | hCoV-19/Wales/PHWC-49403A/2020\|EPI_ISL_726057\|2020 | ··········K········· | ···················· | ···················· | ···················· | ···················· | NNNNNNNNNNNNNNNNNNNN | 1 |
| Y | 5392 | hCoV-19/Wales/PHWC-494119/2020\|EPI_ISL_726070\|2020 | ···················· | ···················· | ···················· | ···················· | NNNNNNNNNNNNNNNNNNNN | ···················· | 1 |
| Y | 5394 | hCoV-19/Wales/PHWC-494137/2020\|EPI_ISL_726072\|2020 | ···················· | ···················· | ···················· | ···················· | NNNNNNNNNNNNNNNNNNNN | ···················· | 1 |
| Y | 5399 | hCoV-19/Wales/PHWC-4941FB/2020\|EPI_ISL_726083\|2020 | ···················· | ···················· | ···················· | ···················· | NNNNNNNNNNNNNNNNNNNN | ···················· | 1 |
| Y | 5408 | hCoV-19/Wales/PHWC-4942AD/2020\|EPI_ISL_726094\|2020 | ···················· | ···················· | ···················· | ···················· | NNNNNNNNNNNNNNNNNNNN | NNNNNNNNNNNNNNNNN··· | 1 |
| Y | 5413 | hCoV-19/Wales/PHWC-494304/2020\|EPI_ISL_726099\|2020 | ···················· | ···················· | ···················· | ···················· | ···················· | NNNNNNNNNNNNNNNNNNNN | 1 |
| Y | 5415 | hCoV-19/Wales/PHWC-499967/2020\|EPI_ISL_727381\|2020 | ···················· | ···················· | ···················· | ···················· | NNNNNNNNNNNNNNNNNNNN | NNNNNNNNNNNNNNNNNNNN | 1 |
| Y | 5416 | hCoV-19/Wales/PHWC-499985/2020\|EPI_ISL_727382\|2020 | ···················· | ···················· | ···················· | ···················· | ·······NNNNNNNNNNNNN | NNNNNNNNNNNNNNNNNNNN | 1 |
| Y | 5419 | hCoV-19/Wales/PHWC-4999C1/2020\|EPI_ISL_727385\|2020 | ···················· | ···················· | ···················· | ···················· | ···················· | NNNNNNNNNNNNNNN····· | 1 |
| Y | 5422 | hCoV-19/Wales/PHWC-499A19/2020\|EPI_ISL_727389\|2020 | ···················· | ···················· | ···················· | ···················· | NNN················· | NNNNNNNNNNNNNNNNNNNN | 1 |
| Y | 5424 | hCoV-19/Wales/PHWC-499A37/2020\|EPI_ISL_727391\|2020 | ···················· | ···················· | ···················· | ···················· | ···················· | NNNNNNNNNNNNNNNNNNNN | 1 |
| Y | 5425 | hCoV-19/Wales/PHWC-499A46/2020\|EPI_ISL_727392\|2020 | ···················· | ·····A·············· | ···················· | ···················· | NNNNNNNNNNNNNNNNNNNN | NNNNNNNNNNNNNNNNNNNN | 1 |
| Y | 5428 | hCoV-19/Wales/PHWC-499A82/2020\|EPI_ISL_727396\|2020 | ···················· | ···················· | ···················· | ···················· | NNNNNNNNNNNNNNNNNNNN | NNNNNNNNNNNNNNNNNNNN | 1 |
| Y | 5429 | hCoV-19/Wales/PHWC-499A91/2020\|EPI_ISL_727397\|2020 | ···················· | ···················· | ···················· | ···················· | NNNNNNNNNNNNNNNNNNNN | NNNNNNNNNNNNNNNNNNNN | 1 |
| Y | 5444 | hCoV-19/Russia/AST-RII-MH4474S/2020\|EPI_ISL_733207 | ···················· | ···················· | ···················· | ···················· | NNNNNNNNNNNNNNNNNNNN | ··············NNNNNN | 1 |
| Y | 5459 | hCoV-19/Russia/SVE-RII-MH4693S/2020\|EPI_ISL_733238 | ···················· | ···················· | ···················· | ···················· | ··N················· | ··············NNNNNN | 1 |
| Y | 5478 | hCoV-19/Russia/KDA-RII-MH4894S/2020\|EPI_ISL_733281 | ···················· | ···················· | ···················· | ···················· | NNNNNNNNNNNNNNNNNNNN | ···················· | 1 |
| Y | 5479 | hCoV-19/Russia/KDA-RII-MH4917S/2020\|EPI_ISL_733284 | ···················· | ···················· | ···················· | ···················· | NNNNNNNNNNNNNN······ | ···················· | 1 |
| Y | 5482 | hCoV-19/Russia/KDA-RII-MH4938S/2020\|EPI_ISL_733290 | ···················· | ···················· | ···················· | ···················· | NNNNNNNNNNNNNNNNNNNN | ···················· | 1 |
| Y | 5636 | hCoV-19/Wales/PHWC-494340/2020\|EPI_ISL_726103\|2020 | ···················· | ···················· | ···················· | ···················· | NNNNNNNNNNNNNNNNNNNN | ···················· | 1 |
| Y | 5644 | hCoV-19/Wales/PHWC-4943F5/2020\|EPI_ISL_726111\|2020 | ···················· | ···················· | ···················· | ···················· | NNNNNNNNNNNNNNNNNNNN | NNNNNNNNNNNNNNN····· | 1 |
| Y | 5648 | hCoV-19/Wales/PHWC-49445C/2020\|EPI_ISL_726117\|2020 | ···················· | ···················· | ···················· | ···················· | NNNNNNNNNNNNNNNNNNNN | ···················· | 1 |
| Y | 5652 | hCoV-19/Wales/PHWC-494498/2020\|EPI_ISL_726121\|2020 | ···················· | ···················· | ···················· | ···················· | NNNNNNNNNNNNNNNNNNN· | ···················· | 1 |
| Y | 5656 | hCoV-19/Wales/PHWC-4944D4/2020\|EPI_ISL_726125\|2020 | ···················· | ···················· | ···················· | ···················· | NNNNNNNNNNNNNNNNNNNN | NNNNNNNNNNNNNNNNNNNN | 1 |
| Y | 5658 | hCoV-19/Wales/PHWC-4944F2/2020\|EPI_ISL_726127\|2020 | ···················· | ···················· | ···················· | ···················· | NNNNNNNNNNNNNN······ | ···················· | 1 |
| Y | 5666 | hCoV-19/Wales/PHWC-494595/2020\|EPI_ISL_726136\|2020 | ···················· | ···················· | ···················· | ···················· | NNNNNNNNNNNNNNNNNNNN | ···················· | 1 |
| Y | 5668 | hCoV-19/Wales/PHWC-4945C2/2020\|EPI_ISL_726139\|2020 | ···················· | ···················· | ···················· | ···················· | ···················· | NNNNN·······NNN····· | 1 |
| Y | 5671 | hCoV-19/Wales/PHWC-4945FF/2020\|EPI_ISL_726142\|2020 | ···················· | ···················· | ···················· | ···················· | NNNNNNNNNNNNNNNNNNNN | ···················· | 1 |
| Y | 5673 | hCoV-19/Wales/PHWC-49461A/2020\|EPI_ISL_726144\|2020 | ···················· | ···················· | ···················· | ···················· | NNNNNNNNNNNNNNNNNNNN | NNNNNNNNNNNNNNNNNNNN | 1 |
| Y | 5682 | hCoV-19/Wales/PHWC-4946A1/2020\|EPI_ISL_726153\|2020 | ···················· | ···················· | ···················· | ···················· | NNNNNNNNNNNNNNNNNNNN | ···················· | 1 |
| Y | 5700 | hCoV-19/Wales/PHWC-49494E/2020\|EPI_ISL_726191\|2020 | ···················· | ···················· | ···················· | ···················· | NNNNNNNNNNNNNNNNNNNN | NNNNNNNNNNNNNNNNNN·· | 1 |
| Y | 5702 | hCoV-19/Wales/PHWC-49496C/2020\|EPI_ISL_726193\|2020 | ···················· | ···················· | ···················· | ···················· | NNNNNNNNNNNNNNNNNNNN | NNNNNNNNNNNNNNN····· | 1 |
| Y | 5707 | hCoV-19/Wales/PHWC-499ADD/2020\|EPI_ISL_727400\|2020 | ···················· | ···················· | ···················· | ···················· | NNNNNNNNNNNNNNNNNNNN | NNNNNNNNNNNNNNNNNNNN | 1 |
| Y | 5709 | hCoV-19/Wales/PHWC-499B16/2020\|EPI_ISL_727403\|2020 | ···················· | ···················· | ···················· | ···················· | NNNNNNNNNNNNNNNNNNNN | NNNNNNNNNNNNNNNNNNNN | 1 |
| Y | 5710 | hCoV-19/Wales/PHWC-499B25/2020\|EPI_ISL_727404\|2020 | ···················· | ···················· | ···················· | ···················· | NNNNNNNNNNNNNNNNNNNN | NNNNNNNNNNNNNNNNNNNN | 1 |
| Y | 5711 | hCoV-19/Wales/PHWC-499B34/2020\|EPI_ISL_727405\|2020 | ···················· | ···················· | ···················· | ···················· | NNNNNNNNNNNNNNNNNNNN | NNNNNNNNNNNNNNNNNNNN | 1 |
| Y | 5714 | hCoV-19/Wales/PHWC-499B70/2020\|EPI_ISL_727408\|2020 | ···················· | ···················· | ···················· | ···················· | ·······NNNNNNNNNNNNN | NNNNNNN············· | 1 |
| Y | 5715 | hCoV-19/Wales/PHWC-499B9E/2020\|EPI_ISL_727410\|2020 | ···················· | ···················· | ···················· | ···················· | NNNNNNNNNNNNNNNNNNNN | NNNNNNNNNNNNNNNNNNNN | 1 |
| Y | 5717 | hCoV-19/Wales/PHWC-499BCB/2020\|EPI_ISL_727413\|2020 | ·T·················· | ···················· | ···················· | ···················· | NNNNNNNNNNNNNNNNNNNN | ···················· | 1 |
| Y | 5718 | hCoV-19/Wales/PHWC-499BE9/2020\|EPI_ISL_727415\|2020 | ···················· | ···················· | ···················· | ···················· | NNNNNNNNNNNNNNNNNNNN | ···················· | 1 |
| Y | 5721 | hCoV-19/Wales/PHWC-499C13/2020\|EPI_ISL_727418\|2020 | ···················· | ···················· | ···················· | ···················· | NNNNNNNNNNNNNNNNNNNN | NNNNNNNNNNNNNNNNNNNN | 1 |
| Y | 5726 | hCoV-19/Wales/PHWC-499C7D/2020\|EPI_ISL_727424\|2020 | ···················· | ···················· | ···················· | ···················· | NNNNNNNNNNNNNNNNNNNN | NNNNNNNNNNNNNNNNNNNN | 1 |
| Y | 5727 | hCoV-19/Wales/PHWC-499C9B/2020\|EPI_ISL_727426\|2020 | ···················· | ···················· | ···················· | ···················· | NNNNNNNNNNNNNNNNNNNN | NNNNNNNNNNNNNNNNNNNN | 1 |
| Y | 5731 | hCoV-19/Wales/PHWC-499CE6/2020\|EPI_ISL_727431\|2020 | ···················· | ···················· | ···················· | ···················· | NNNNNNNNNNNNNNNNNNNN | NNNNNNNNNNNNNNNNNNNN | 1 |
| Y | 5732 | hCoV-19/Wales/PHWC-499D01/2020\|EPI_ISL_727433\|2020 | ···················· | ···················· | ···················· | ···················· | ···················· | NNNNNNNNNNN········· | 1 |

| | | | | | | | | | |
|---|---|---|---|---|---|---|---|---|---|
| Y | 5735 | hCoV-19/Wales/PHWC-499D6B/2020\|EPI_ISL_727439\|2020 | ···················· | ···················· | ···················· | ···················· | NNNNNNNNNNNNNNNNNNNN | NNNNNNNNNNNNNNNNNNNN | 1 |
| Y | 5739 | hCoV-19/Wales/PHWC-499DA7/2020\|EPI_ISL_727443\|2020 | ···················· | ···················· | ···················· | ···················· | NNNNNNNNNNNNNNNNNNNN | NNNNNNNNNNNNNNNNNNNN | 1 |
| Y | 5744 | hCoV-19/Wales/PHWC-499E1D/2020\|EPI_ISL_727450\|2020 | ···················· | ···················· | ···················· | ···················· | NNNNNNNNNNNNNNNNNNNN | NNNNNNNNNNNNNNN····· | 1 |
| Y | 5745 | hCoV-19/Wales/PHWC-499E2C/2020\|EPI_ISL_727451\|2020 | ···················· | ···················· | ···················· | ···················· | NNNNNNNNNNNNNNNNNNNN | NNNNNNNNNNNNNNNNNNNN | 1 |
| Y | 5746 | hCoV-19/Wales/PHWC-499E3B/2020\|EPI_ISL_727452\|2020 | ···················· | ···················· | ···················· | ···················· | NNNNNNNNNNNNNNNNNN·· | NN·················· | 1 |
| Y | 5751 | hCoV-19/Wales/PHWC-499EA4/2020\|EPI_ISL_727459\|2020 | ···················· | ···················· | ···················· | ···················· | NNNNNNNNNNNNNNNNNNNN | NNNNNNNNNNNNNNNNNNNN | 1 |
| Y | 5752 | hCoV-19/Wales/PHWC-499EB3/2020\|EPI_ISL_727460\|2020 | ···················· | ···················· | ···················· | ···················· | NNNNNNNNNNNNNNNNNNNN | NNNNNNNNNNNNNNNNNNNN | 1 |
| Y | 5754 | hCoV-19/Wales/PHWC-499ED1/2020\|EPI_ISL_727462\|2020 | ···················· | ···················· | ···················· | ···················· | NNNNNNNNNNNNNNNNNNNN | NNNNNNNNNNNNNNNNNNNN | 1 |
| Y | 5755 | hCoV-19/Wales/PHWC-499EE0/2020\|EPI_ISL_727463\|2020 | ···················· | ···················· | ···················· | ···················· | NNNNNNNNNNNNNNNNNNNN | NNNNN·········N····· | 1 |
| Y | 5756 | hCoV-19/Wales/PHWC-499EFF/2020\|EPI_ISL_727464\|2020 | ···················· | ···················· | ···················· | ···················· | NNNNNNNNNNNNNNNNNNNN | ···················· | 1 |
| Y | 5760 | hCoV-19/Wales/PHWC-499F38/2020\|EPI_ISL_727468\|2020 | ···················· | ···················· | ···················· | ···················· | NNNNNNNNNNNNNNNNNNNN | NNNNNNNNNNNNNNNNNNNN | 1 |
| Y | 5761 | hCoV-19/Wales/PHWC-499F47/2020\|EPI_ISL_727469\|2020 | ···················· | ···················· | ···················· | ···················· | NNN··········N······ | ···················· | 1 |
| Y | 5762 | hCoV-19/Wales/PHWC-499F83/2020\|EPI_ISL_727473\|2020 | ···················· | ···················· | ···················· | ···················· | NNNNNNNNNNNNNNNNNNNN | ···················· | 1 |
| Y | 5764 | hCoV-19/Wales/PHWC-499FB0/2020\|EPI_ISL_727476\|2020 | ···················· | ···················· | ···················· | ···················· | NNNNNNNNNNNNNNNNNNNN | NNNNNNNNNNNNNNNNNNNN | 1 |
| Y | 5766 | hCoV-19/Wales/PHWC-499FDE/2020\|EPI_ISL_727478\|2020 | ···················· | ···················· | ···················· | ···················· | NNNNNNNNNNNNNNNNNNNN | NNNNNNNNNNNNNNNN···· | 1 |
| Y | 5767 | hCoV-19/Wales/PHWC-499FED/2020\|EPI_ISL_727479\|2020 | ···················· | ···················· | ···················· | ···················· | NNNNNNNNNNNNNNNNNNNN | NNNNNNNNNNNNNNNNNNNN | 1 |
| Y | 5770 | hCoV-19/Wales/PHWC-49A016/2020\|EPI_ISL_727482\|2020 | ···················· | ···················· | ···················· | ···················· | NNNNNNNNNNNNNNNNNNNN | NNNNNNNNNNNNNNNNNNNN | 1 |
| Y | 6148 | hCoV-19/Wales/PHWC-494A2D/2020\|EPI_ISL_726204\|2020 | ···················· | ···················· | ···················· | ···················· | NNNNNNNNNNNNNNNNNNNN | NNNNNNNNNNNNNNNNNNNN | 1 |
| Y | 6150 | hCoV-19/Wales/PHWC-494A4B/2020\|EPI_ISL_726206\|2020 | ···················· | ···················· | ···················· | ···················· | NNNNNNNNNNNNNNNNNNNN | NNNNNNNNNNNNNNNNNNNN | 1 |
| Y | 6153 | hCoV-19/Wales/PHWC-494A87/2020\|EPI_ISL_726210\|2020 | ···················· | ···················· | ···················· | ···················· | NNNNNNNNNNNNNNNNNNNN | NN·················· | 1 |
| Y | 6154 | hCoV-19/Wales/PHWC-494A96/2020\|EPI_ISL_726211\|2020 | ···················· | ···················· | ···················· | ···················· | NNNNNNNNNNNNNNNNNNNN | ·····NNNNNNNNNNNNNNN | 1 |
| Y | 6161 | hCoV-19/Wales/PHWC-494B2A/2020\|EPI_ISL_726219\|2020 | ···················· | ···················· | ···················· | ···················· | NNNNNNNNNNNNNNNNNNNN | ···················· | 1 |
| Y | 6163 | hCoV-19/Wales/PHWC-494B66/2020\|EPI_ISL_726222\|2020 | ···················· | ···················· | ···················· | ···················· | NNNNNNNNNNNNNNNNNNNN | NNNNNNNNNNNNNNNNNNNN | 1 |
| Y | 6164 | hCoV-19/Wales/PHWC-494B84/2020\|EPI_ISL_726224\|2020 | ···················· | ···················· | ···················· | ···················· | NNNNNNNNNNNNNNNNNNNN | NNNNNNNNNNNNNNNNNNNN | 1 |
| Y | 6165 | hCoV-19/Wales/PHWC-494B93/2020\|EPI_ISL_726225\|2020 | ···················· | ···················· | ···················· | ···················· | NNNNNNNNNNNNNNNNNNNN | NNNNNNNNNNNNNNNNNNNN | 1 |
| Y | 6167 | hCoV-19/Wales/PHWC-494BC0/2020\|EPI_ISL_726227\|2020 | ···················· | ···················· | ···················· | ···················· | NNNNNNNNNNNNNNNNNNNN | ··············NNNNNN | 1 |
| Y | 6174 | hCoV-19/Wales/PHWC-494C36/2020\|EPI_ISL_726234\|2020 | ···················· | ···················· | ···················· | ···················· | NNN················· | NNNNNNNN············ | 1 |
| Y | 6182 | hCoV-19/Wales/PHWC-494CFA/2020\|EPI_ISL_726242\|2020 | ···················· | ···················· | ···················· | ···················· | NNNNNNNNNNNNNNNNNNNN | NNNNNNNNNNNNNNNNNNNN | 1 |
| Y | 6183 | hCoV-19/Wales/PHWC-494D15/2020\|EPI_ISL_726244\|2020 | ···················· | ···················· | ···················· | ···················· | NNNNNNNNNNNNNNNNNNNN | NNNNNNNNNNNNNNNNNNNN | 1 |
| Y | 6184 | hCoV-19/Wales/PHWC-494D24/2020\|EPI_ISL_726245\|2020 | ···················· | ···················· | ···················· | ···················· | NNNNNNNNNNNNNNNNNNNN | NNNNNNNNNNNNNNNNNNNN | 1 |
| Y | 6188 | hCoV-19/Wales/PHWC-494D60/2020\|EPI_ISL_726249\|2020 | ···················· | ···················· | ···················· | ···················· | ···················· | NNNNNNNN············ | 1 |
| Y | 6189 | hCoV-19/Wales/PHWC-494D9D/2020\|EPI_ISL_726252\|2020 | ···················· | ···················· | ···················· | ···················· | NNNNNNNNNNNNNNNNNNNN | NNNNNNNNNNNNNNNNNNNN | 1 |
| Y | 6193 | hCoV-19/Wales/PHWC-494EF4/2020\|EPI_ISL_726270\|2020 | ···················· | ···················· | ···················· | ···················· | ·················NNN | ········NNNNNNN····· | 1 |
| Y | 6195 | hCoV-19/Wales/PHWC-494F3D/2020\|EPI_ISL_726273\|2020 | ···················· | ···················· | ···················· | ···················· | NNNNNNNNNNNNNNNNNNNN | ···················· | 1 |
| Y | 6198 | hCoV-19/Wales/PHWC-494F6A/2020\|EPI_ISL_726276\|2020 | ···················· | ···················· | ···················· | ···················· | NNNNNNNNNNNNNNNNNNNN | ···················· | 1 |
| Y | 6199 | hCoV-19/Wales/PHWC-494FA6/2020\|EPI_ISL_726280\|2020 | ···················· | ···················· | ···················· | ···················· | ···················· | NNNNNNNNNNNNNNNNNNNN | 1 |
| Y | 6200 | hCoV-19/Wales/PHWC-494FB5/2020\|EPI_ISL_726281\|2020 | ···················· | ···················· | ···················· | ···················· | ···················· | NNNNN··············· | 1 |
| Y | 6201 | hCoV-19/Wales/PHWC-494FC4/2020\|EPI_ISL_726282\|2020 | ···················· | ···················· | ···················· | ···················· | NNNNNNNNNNNNNNNNNNNN | ···················· | 1 |
| Y | 6203 | hCoV-19/Wales/PHWC-494FF1/2020\|EPI_ISL_726285\|2020 | ···················· | ···················· | ···················· | ···················· | NNNNNNNNNNNNNNNNNNNN | NNNNNNNNNNNNNNNNNNNN | 1 |
| Y | 6206 | hCoV-19/Wales/PHWC-49502A/2020\|EPI_ISL_726288\|2020 | ···················· | ···················· | ···················· | ···················· | NNNNNNNNNNNNNNNNNNNN | ···················· | 1 |
| Y | 6207 | hCoV-19/Wales/PHWC-495039/2020\|EPI_ISL_726289\|2020 | ···················· | ···················· | ···················· | ···················· | NNNNNNNNNNNN········ | ···················· | 1 |
| Y | 6208 | hCoV-19/Wales/PHWC-49A140/2020\|EPI_ISL_727500\|2020 | ···················· | ···················· | ···················· | ···················· | NNNNNNNNNNNNNNNNNNNN | ················NNNN | 1 |
| Y | 6210 | hCoV-19/Wales/PHWC-49A17D/2020\|EPI_ISL_727502\|2020 | ···················· | ···················· | ···················· | ···················· | NNNNNNNNNNNNNNNNNNNN | NNNNNNNNNNNNNNNNNNNN | 1 |
| Y | 6219 | hCoV-19/Wales/PHWC-49A210/2020\|EPI_ISL_727512\|2020 | ···················· | ···················· | ···················· | ···················· | NNNNNNNNNNNNNNNNNNNN | NNNNNNNNNNNNNNNNNNNN | 1 |
| Y | 6220 | hCoV-19/Wales/PHWC-49A22F/2020\|EPI_ISL_727513\|2020 | ···················· | ·····A·············· | ···················· | ···················· | ···················· | NNNNNNN············· | 1 |
| Y | 6221 | hCoV-19/Wales/PHWC-49A23E/2020\|EPI_ISL_727514\|2020 | ···················· | ···················· | ···················· | ···················· | ···················· | NNNNNNNNNNNNNNNNNNNN | 1 |
| Y | 6222 | hCoV-19/Wales/PHWC-49A24D/2020\|EPI_ISL_727515\|2020 | ···················· | ···················· | ···················· | ···················· | NNNNNNNNNNNNNNNNNNNN | ···················· | 1 |
| Y | 6223 | hCoV-19/Wales/PHWC-49A25C/2020\|EPI_ISL_727516\|2020 | ···················· | ···················· | ···················· | ···················· | NNNNNNNNNNNNNNNNNNNN | NNNNNNNNNNNNNNNNNNNN | 1 |
| Y | 6224 | hCoV-19/Wales/PHWC-49A26B/2020\|EPI_ISL_727517\|2020 | ···················· | ···················· | ···················· | ···················· | NNNNNNNNNNNNNNNNNNNN | NNNNNNNNNNNNNNNNNNNN | 1 |
| Y | 6225 | hCoV-19/Wales/PHWC-49A27A/2020\|EPI_ISL_727518\|2020 | ···················· | ···················· | ···················· | ···················· | NNNNNNNNNNNNNNNNNNNN | NNNNNNNNNNNNNNNNNNNN | 1 |
| Y | 6228 | hCoV-19/Wales/PHWC-49A2A7/2020\|EPI_ISL_727521\|2020 | ···················· | ···················· | ···················· | ···················· | NNNNNNNNNNNNNNNNNNNN | NNNNNNNNNNNNNNNNNNNN | 1 |
| Y | 6229 | hCoV-19/Wales/PHWC-49A2B6/2020\|EPI_ISL_727522\|2020 | ···················· | ···················· | ···················· | ···················· | NNNNNNNNNNNNNNNNNNNN | NNNNNNNNNNNNNNNNNNNN | 1 |
| Y | 6230 | hCoV-19/Wales/PHWC-49A2C5/2020\|EPI_ISL_727523\|2020 | ···················· | ···················· | ···················· | ···················· | NNNNNNNNNNNNNNNNNNNN | NNNNNNNNNNNNNNNNNNNN | 1 |
| Y | 6234 | hCoV-19/Wales/PHWC-49A31D/2020\|EPI_ISL_727528\|2020 | ···················· | ···················· | ···················· | ···················· | ···················· | NNNNNNNNNNNNNNN····· | 1 |
| Y | 6237 | hCoV-19/Wales/PHWC-49A34A/2020\|EPI_ISL_727531\|2020 | ···················· | ···················· | ···················· | ···················· | NNNNNNNNNNNNNNNNNNNN | NNNNNNNNNNNNNNNNNNNN | 1 |
| Y | 6238 | hCoV-19/Wales/PHWC-49A359/2020\|EPI_ISL_727532\|2020 | ···················· | ···················· | ···················· | ···················· | ···················· | NNNNNNNNNNNNNNNNNNNN | 1 |
| Y | 6239 | hCoV-19/Wales/PHWC-49A368/2020\|EPI_ISL_727533\|2020 | ···················· | ···················· | ···················· | ···················· | NNNNNNNNNNNNNNNNNNNN | NNNNNNNNNNNNNNNNNNNN | 1 |
| Y | 6242 | hCoV-19/Wales/PHWC-49A3A4/2020\|EPI_ISL_727536\|2020 | ···················· | ···················· | ···················· | ···················· | NNNNNNN······NNNNNNN | NNNNNNNNNNNNNNNNNNNN | 1 |
| Y | 6251 | hCoV-19/Wales/PHWC-49A438/2020\|EPI_ISL_727545\|2020 | ···················· | ···················· | ···················· | ···················· | NNNNNNNNNNNNNNNNNNNN | NNNNNNNNNNNNNNNNNNNN | 1 |
| Y | 6254 | hCoV-19/Wales/PHWC-49A474/2020\|EPI_ISL_727549\|2020 | ···················· | ···················· | ···················· | ···················· | NNNNNNNNNNNNNNNNNNNN | NNNNNNNNNNNNNNNNNNNN | 1 |
| Y | 6283 | hCoV-19/Wales/PHWC-49A6BA/2020\|EPI_ISL_727582\|2020 | ···················· | ···················· | ···················· | ···················· | NNNNNNNNNNNNNNNNNNNN | NNNNNNNNNNNNNNNNNNNN | 1 |
| Y | 6290 | hCoV-19/Wales/PHWC-49A73F/2020\|EPI_ISL_727589\|2020 | ···················· | ···················· | ···················· | ···················· | NNNNNNNNNNNNNNNNNNNN | NNNNNNNNNNNNNNNNNNNN | 1 |
| Y | 6402 | hCoV-19/Wales/PHWC-494E30/2020\|EPI_ISL_726260\|2020 | ···················· | ···················· | ···················· | ···················· | NNNNNNNNNNNNNNNNNNNN | NNNNNNNNNNNNNNNNNNNN | 1 |
| Y | 6403 | hCoV-19/Wales/PHWC-494E4F/2020\|EPI_ISL_726261\|2020 | ···················· | ···················· | ···················· | ···················· | NNNNNNNNNNNNNNNNNNNN | NNNNNNNNNNNNNNNNNNNN | 1 |
| Y | 6404 | hCoV-19/Wales/PHWC-494E5E/2020\|EPI_ISL_726262\|2020 | ···················· | ···················· | ···················· | ···················· | NNNNNNNNNNNNNNNNNNNN | NNNNNNNNNNNNNNNNNNNN | 1 |
| Y | 6408 | hCoV-19/Wales/PHWC-494EC7/2020\|EPI_ISL_726268\|2020 | ···················· | ···················· | ···················· | ···················· | NNNNNNNNNNNNNNNNNNNN | NNNNNNNNNNNNNNNNNNNN | 1 |
| Y | 6409 | hCoV-19/Wales/PHWC-495048/2020\|EPI_ISL_726290\|2020 | ···················· | ···················· | ···················· | ···················· | NNNNNNNNNNNNNNNNNNNN | NNNNNNNNNNNNNNNNNNNN | 1 |
| Y | 6410 | hCoV-19/Wales/PHWC-495057/2020\|EPI_ISL_726291\|2020 | ···················· | ···················· | ···················· | ···················· | NNNNNNNNNNNNNNNNNNNN | NNNNNNNNNNNNNNN····· | 1 |
| Y | 6411 | hCoV-19/Wales/PHWC-495075/2020\|EPI_ISL_726293\|2020 | ···················· | ···················· | ···················· | ···················· | NNNNNNNNNNNNNNNNNNNN | NNNNNNNNNNNNNNNNNNNN | 1 |
| Y | 6412 | hCoV-19/Wales/PHWC-495084/2020\|EPI_ISL_726294\|2020 | ···················· | ···················· | ···················· | ···················· | NNNNNNNNNNNNNNNNNNNN | NNNNNNNNNNNNNNNNNNNN | 1 |
| Y | 6414 | hCoV-19/Wales/PHWC-4950A2/2020\|EPI_ISL_726296\|2020 | ···················· | ···················· | ···················· | ···················· | NNNNNNNNNNNNNNNNNNNN | N··········NNNN····· | 1 |
| Y | 6425 | hCoV-19/Wales/PHWC-49A7A8/2020\|EPI_ISL_727596\|2020 | ···················· | ···················· | ···················· | ···················· | NNNNNNNNNNNNNNNNNNNN | NNNNNNNNNNNNNNNNNNNN | 1 |
| Y | 6431 | hCoV-19/Wales/PHWC-49A81E/2020\|EPI_ISL_727603\|2020 | ···················· | ···················· | ···················· | ···················· | ··NNNNNNNNNNNNNNNNNN | NNNNNNNNNNNNNNN····· | 1 |
| Y | 6462 | hCoV-19/Portugal/PT1905/2020\|EPI_ISL_732119\|2020-1 | ···················· | ···················· | ···················· | ···················· | NNNNNNNNNNNNNNNNNNNN | ··N··········N······ | 1 |
| Y | 6478 | hCoV-19/Portugal/PT1923/2020\|EPI_ISL_732135\|2020-1 | ···················· | ···················· | ···················· | ···················· | NNNNNNNNNNNNNNNNNNNN | ·NNNNNNNNN······N··N | 1 |
| Y | 6516 | hCoV-19/Portugal/PT1974/2020\|EPI_ISL_732198\|2020-1 | ···················· | ···················· | ···················· | ···················· | NNNNNNNNNNNNNNNNNNNN | ···················· | 1 |
| Y | 6519 | hCoV-19/Russia/SVE-RII-MH9083S/2020\|EPI_ISL_733407 | ···················· | ···················· | ···················· | NNNNNNNNNNNNNNNNNNNN | NNNNNNNNNN·········· | ···················· | 1 |
| Y | 6675 | hCoV-19/England/NORW-21F594/2020\|EPI_ISL_725029\|20 | ···················· | ···················· | ···················· | ···················· | NNNNNNNNNNNNNNNNNNNN | NNNNNNNNNNNNNNNNNNNN | 1 |
| Y | 6681 | hCoV-19/Wales/PHWC-4950EE/2020\|EPI_ISL_726300\|2020 | ···················· | ···················· | ···················· | ···················· | NNNNNNNNNNNNNNNNNNNN | ················NNNN | 1 |
| Y | 6682 | hCoV-19/Wales/PHWC-4950FD/2020\|EPI_ISL_726301\|2020 | ···················· | ···················· | ···················· | ···················· | NNNNNNNNNNNNNNNNNNNN | NNNNNNNNNNNNNNNNNNNN | 1 |

| | | | | | | | | |
|---|---|---|---|---|---|---|---|---|
| Y | 6684 hCoV-19/Wales/PHWC-495127/2020\|EPI_ISL_726303\|2020 | ···················· | ···················· | ···················· | ···················· | NNNNNNNNNNNNNNNNNNNN | NNNNNNNNNNNNNNNNNNNN | 1 |
| Y | 6689 hCoV-19/Wales/PHWC-4951BE/2020\|EPI_ISL_726309\|2020 | ···················· | ···················· | ···················· | ···················· | NNNNNNNNNNNNNNNNNNNN | NNNNNNNNNNNNNNNNNNNN | 1 |
| Y | 6690 hCoV-19/Wales/PHWC-4951DC/2020\|EPI_ISL_726310\|2020 | ···················· | ···················· | ···················· | ···················· | NNNNNNNNNNNNNNNNNNNN | NNNNNNNNNNNNNNNNNNNN | 1 |
| Y | 6693 hCoV-19/Wales/PHWC-495260/2020\|EPI_ISL_726318\|2020 | ···················· | ···················· | ···················· | ···················· | NNNNNNNNNNNNNNNNNNNN | NNNNNNNNNNNNNNNNNNNN | 1 |
| Y | 6694 hCoV-19/Wales/PHWC-49527F/2020\|EPI_ISL_726319\|2020 | ···················· | ···················· | ···················· | ···················· | ···················· | NNNNNNNNNNNNNNNNNNNN | 1 |
| Y | 6696 hCoV-19/Wales/PHWC-4952BB/2020\|EPI_ISL_726322\|2020 | ···················· | ···················· | ···················· | ···················· | NNNNNNNNNNNNNNNNNNNN | NNNNNNNNNNNNNNNNNNNN | 1 |
| Y | 6698 hCoV-19/Wales/PHWC-4952E8/2020\|EPI_ISL_726325\|2020 | ···················· | ···················· | ···················· | ···················· | NNNNNNNNNNNNNNNNNNNN | NNNNNNNNNNNNNNN····· | 1 |
| Y | 6699 hCoV-19/Wales/PHWC-4952F7/2020\|EPI_ISL_726326\|2020 | ···················· | ···················· | ···················· | ···················· | NNNNNNNNNNNNNNNNNNNN | NNNNNNNNNNNNNNNNNNNN | 1 |
| Y | 6700 hCoV-19/Wales/PHWC-495303/2020\|EPI_ISL_726327\|2020 | ···················· | ···················· | ···················· | ···················· | NNNNNNNNNNNNNNNNNNNN | NNNNNNNNNNNNNNNNNNNN | 1 |
| Y | 6701 hCoV-19/Wales/PHWC-495312/2020\|EPI_ISL_726328\|2020 | ···················· | ···················· | ···················· | ···················· | NNNNNNNNNNNNNNNNNNNN | NNNNNNNNNNNNNNNNNNNN | 1 |
| Y | 6702 hCoV-19/Wales/PHWC-495321/2020\|EPI_ISL_726329\|2020 | ···················· | ···················· | ···················· | ···················· | NNNNNNNNNNNNNNNNNNNN | NNNNNNNNNNNNNNNN···· | 1 |
| Y | 6703 hCoV-19/Wales/PHWC-495330/2020\|EPI_ISL_726330\|2020 | ···················· | ···················· | ···················· | ···················· | NNNNNNNNNNNNNNNNNNNN | NNNNNNNNNNNNNNNNNNNN | 1 |
| Y | 6704 hCoV-19/Wales/PHWC-49535E/2020\|EPI_ISL_726332\|2020 | ···················· | ···················· | ···················· | ···················· | NNNNNNNNNNNNNNNNNNNN | NNNNNNNNNNNNNNNNNNNN | 1 |
| Y | 6705 hCoV-19/Wales/PHWC-49536D/2020\|EPI_ISL_726333\|2020 | ···················· | ·····A·············· | ···················· | ···················· | NNNNNNNNNNNNNNNNNNNN | NNNNNNNNNNNNNNNNNNNN | 1 |
| Y | 6706 hCoV-19/Wales/PHWC-49538B/2020\|EPI_ISL_726335\|2020 | ···················· | ···················· | ···················· | ···················· | NNNNNNNNNNNNNNNNNNNN | NNNNNNNNNNNNNNNNNNNN | 1 |
| Y | 6708 hCoV-19/Wales/PHWC-4953B8/2020\|EPI_ISL_726338\|2020 | ···················· | ···················· | ···················· | ···················· | NNNNNNNNNNNNNNNNNNNN | NNNNNNNNNNNNNNNNNNNN | 1 |
| Y | 6709 hCoV-19/Wales/PHWC-4953C7/2020\|EPI_ISL_726339\|2020 | ···················· | ···················· | ···················· | ···················· | ···················· | NNNNNNNNNNNNNNNNNNNN | 1 |
| Y | 6710 hCoV-19/Wales/PHWC-4953D6/2020\|EPI_ISL_726340\|2020 | ···················· | ···················· | ···················· | ···················· | NNNNNNNNNNNNNNNNNNNN | NNNNNNNNNNNNNNNNN··· | 1 |
| Y | 6712 hCoV-19/Wales/PHWC-495400/2020\|EPI_ISL_726343\|2020 | ···················· | ···················· | ···················· | ···················· | NNNNNNNNNNNNNNNNNNNN | NNNNNNNNNNNNNNNNNNNN | 1 |
| Y | 6713 hCoV-19/Wales/PHWC-49541F/2020\|EPI_ISL_726344\|2020 | ···················· | ···················· | ···················· | ···················· | NNNNNNNNNNNNNNNNNNNN | NNNNNNNNNNNNNNNNNNNN | 1 |
| Y | 6714 hCoV-19/Wales/PHWC-49543D/2020\|EPI_ISL_726346\|2020 | ···················· | ···················· | ···················· | ···················· | NNNNNNNNNNNNNNNNNNNN | NNNNNNNNNNNNNNNNNNNN | 1 |
| Y | 6718 hCoV-19/Wales/PHWC-495497/2020\|EPI_ISL_726351\|2020 | ···················· | ···················· | ···················· | ···················· | NNNNNNNNNNNNNNNNNNNN | ···················· | 1 |
| Y | 6721 hCoV-19/Wales/PHWC-4954C4/2020\|EPI_ISL_726354\|2020 | ···················· | ···················· | ···················· | ···················· | NNNNNNNNNNNNNNNNNNNN | NNNNNNNNNNNNNNNNNNNN | 1 |
| Y | 6724 hCoV-19/Wales/PHWC-49551C/2020\|EPI_ISL_726357\|2020 | ···················· | ···················· | ···················· | ···················· | NNNNNNNNNNNNNNNNNNNN | ···················· | 1 |
| Y | 6725 hCoV-19/Wales/PHWC-49552B/2020\|EPI_ISL_726358\|2020 | ···················· | ···················· | ···················· | ···················· | NNNNNNNNNNNNNNNNNNNN | NNNNNNNNNNNNNNNNNNNN | 1 |
| Y | 6726 hCoV-19/Wales/PHWC-495549/2020\|EPI_ISL_726360\|2020 | ···················· | ···················· | ···················· | ···················· | NNNNNNNNNNNN········ | ···················· | 1 |
| Y | 6727 hCoV-19/Wales/PHWC-495558/2020\|EPI_ISL_726361\|2020 | ···················· | ···················· | ···················· | ···················· | ···················· | NNNNNNNNNNNNNNN····· | 1 |
| Y | 6731 hCoV-19/Wales/PHWC-4955B2/2020\|EPI_ISL_726367\|2020 | ···················· | ···················· | ···················· | ···················· | NNN················· | NNNNNNNNNNNNNNN····· | 1 |
| Y | 6732 hCoV-19/Wales/PHWC-4955C1/2020\|EPI_ISL_726368\|2020 | ···················· | ···················· | ···················· | ···················· | NNNNNNNNNNNNNNNNNNNN | NNNNNNNNNNNNNNNNNNNN | 1 |
| Y | 6733 hCoV-19/Wales/PHWC-4955D0/2020\|EPI_ISL_726369\|2020 | ···················· | ·····A·············· | ···················· | ···················· | NNNNNNNNNNNNNNNNNNNN | ·····NNNNNNNNNNNNNNN | 1 |
| Y | 6739 hCoV-19/Wales/PHWC-49A975/2020\|EPI_ISL_727623\|2020 | ···················· | ···················· | ···················· | ···················· | NNNNNNNNNNNNNNNNNNNN | ···················· | 1 |
| Y | 6742 hCoV-19/Wales/PHWC-49A9DF/2020\|EPI_ISL_727628\|2020 | ···················· | ···················· | ···················· | ···················· | ······NNNNNNNNNNNNNN | ···················· | 1 |
| Y | 6746 hCoV-19/Wales/PHWC-49AA18/2020\|EPI_ISL_727632\|2020 | ···················· | ···················· | ···················· | ···················· | ···················· | NNNNNNNNNNNNNNN····· | 1 |
| Y | 6752 hCoV-19/Wales/PHWC-49AA81/2020\|EPI_ISL_727639\|2020 | ···················· | ···················· | ···················· | ···················· | NNNNNNNNNNNNNNNNNNNN | NNNNNNNNNNNNNNN····· | 1 |
| Y | 6753 hCoV-19/Wales/PHWC-49AAAF/2020\|EPI_ISL_727641\|2020 | ···················· | ···················· | ···················· | ···················· | NNNNNNNNNNNNNNNNNNNN | NNNNNNNNNNNNNNNNNNNN | 1 |
| Y | 6757 hCoV-19/Wales/PHWC-49AAEB/2020\|EPI_ISL_727645\|2020 | ···················· | ···················· | ···················· | ···················· | NNNNNNNNNNNN········ | NNNNNNNNNNNNNNN···NN | 1 |
| Y | 6765 hCoV-19/Wales/PHWC-49AB8E/2020\|EPI_ISL_727654\|2020 | ···················· | ···················· | ···················· | ···················· | NNNNNNNNNNNNNNNNNNNN | NNNNNNNNNNNNNNNNNNNN | 1 |
| Y | 6766 hCoV-19/Wales/PHWC-49AB9D/2020\|EPI_ISL_727655\|2020 | ···················· | ···················· | ···················· | ···················· | NNNN················ | NNNNNNNNNNNNNNNNNNNN | 1 |
| Y | 6769 hCoV-19/Wales/PHWC-49ABCA/2020\|EPI_ISL_727658\|2020 | ···················· | ···················· | ···················· | ···················· | NNNNNNNNNNNNN······· | NNNNNNNNNNNNNNN····· | 1 |
| Y | 6770 hCoV-19/Wales/PHWC-49ABD9/2020\|EPI_ISL_727659\|2020 | ···················· | ···················· | ···················· | ···················· | ···················· | NNNNNNNNNNNNNNNNNNNN | 1 |
| Y | 6771 hCoV-19/Wales/PHWC-49ABE8/2020\|EPI_ISL_727660\|2020 | ···················· | ···················· | ···················· | ···················· | NNNNNNNNNNNNNNNNNNNN | NNNNNNNNNNNNNNNNNNNN | 1 |
| Y | 6774 hCoV-19/Wales/PHWC-49AC12/2020\|EPI_ISL_727663\|2020 | ···················· | ···················· | ···················· | ···················· | NNNN················ | ···················· | 1 |
| Y | 6775 hCoV-19/Wales/PHWC-49AC21/2020\|EPI_ISL_727664\|2020 | ···················· | ···················· | ···················· | ···················· | NNNNNNNNNNNNNNNNNNNN | NNNNNNNNNNNNNNNNNNNN | 1 |
| Y | 6778 hCoV-19/Wales/PHWC-49AC5E/2020\|EPI_ISL_727667\|2020 | ···················· | ···················· | ···················· | ···················· | NNNNNNNNNNNNNNNNNNNN | NNNNNNNNNNNNNNNNNNNN | 1 |
| Y | 6779 hCoV-19/Wales/PHWC-49AC6D/2020\|EPI_ISL_727668\|2020 | ···················· | ···················· | ···················· | ···················· | NNNNNNNNNNNNNNNNNNNN | NNNNNNNNNNNNNNNNNNNN | 1 |
| Y | 6780 hCoV-19/Wales/PHWC-49ACB8/2020\|EPI_ISL_727672\|2020 | ···················· | ···················· | ···················· | ···················· | NNNNNNNNNNNNNNNNNNNN | NNNNNNNNNNNNNNNNNNNN | 1 |
| Y | 6783 hCoV-19/Wales/PHWC-49AD1F/2020\|EPI_ISL_727677\|2020 | ···················· | ···················· | ···················· | ···················· | NNNNNNNNNNNNNNNNNNNN | NNNNNNNNNNNNNNNNNNNN | 1 |
| Y | 6784 hCoV-19/Wales/PHWC-49AD2E/2020\|EPI_ISL_727678\|2020 | ···················· | ···················· | ···················· | ···················· | NNNNNNNNNNNNNNNNNNNN | NNNNNNNNNNNNNNNNNNNN | 1 |
| Y | 6785 hCoV-19/Wales/PHWC-49AD3D/2020\|EPI_ISL_727679\|2020 | ···················· | ···················· | ···················· | ···················· | NNNNNNNNNNNNNNNNNNNN | NNNNNNNNNNNNNNNNNNNN | 1 |
| Y | 6786 hCoV-19/Wales/PHWC-49AD4C/2020\|EPI_ISL_727680\|2020 | ···················· | ···················· | ···················· | ···················· | NNNNNNNNNNNNNNNNNNNN | NNNNNNNNNNNNNNNNNNNN | 1 |
| Y | 6787 hCoV-19/Wales/PHWC-49AD5B/2020\|EPI_ISL_727681\|2020 | ···················· | ···················· | ···················· | ···················· | NNNNNNNNNNNNNNNNNNNN | NNNNNNNNNNNNNNNNNNNN | 1 |
| Y | 6788 hCoV-19/Wales/PHWC-49AD79/2020\|EPI_ISL_727682\|2020 | ·T·················· | ···················· | ···················· | ···················· | NNNNNNNNNNNNNNNNNNNN | NNNNNNNNNNNNNNNNNNNN | 1 |
| Y | 6790 hCoV-19/Wales/PHWC-49ADB5/2020\|EPI_ISL_727686\|2020 | ···················· | ···················· | ···················· | ···················· | NNNNNNNNNNNNNNNNNNNN | NNNNNNNNNNNNNNNNNNNN | 1 |
| Y | 6791 hCoV-19/Wales/PHWC-49ADC4/2020\|EPI_ISL_727687\|2020 | ···················· | ···················· | ···················· | ···················· | NNNNNNNNNNNNNNNNNNNN | ···················· | 1 |
| Y | 6793 hCoV-19/Wales/PHWC-49ADE2/2020\|EPI_ISL_727689\|2020 | ···················· | ···················· | ···················· | ···················· | ···················· | NNNNNNNNNNNNNNN····· | 1 |
| Y | 6794 hCoV-19/Wales/PHWC-49ADF1/2020\|EPI_ISL_727690\|2020 | ···················· | ···················· | ···················· | ···················· | NNNNNNNNNNNNNNNNNNNN | NNNNNNNNNNNNNNNNNNNN | 1 |
| Y | 6795 hCoV-19/Wales/PHWC-49AE0D/2020\|EPI_ISL_727691\|2020 | ···················· | ···················· | ···················· | ···················· | NNNNNNNNNNNNNNNNNNNN | ···················· | 1 |
| Y | 6796 hCoV-19/Wales/PHWC-49AE1C/2020\|EPI_ISL_727692\|2020 | ···················· | ···················· | ···················· | ···················· | ···················· | NNNNNNNNNNN········· | 1 |
| Y | 6797 hCoV-19/Wales/PHWC-49AE2B/2020\|EPI_ISL_727693\|2020 | ···················· | ···················· | ···················· | ···················· | NNNNNNNNNNNNNNNNNNNN | NNNNNNNNNNNNNNNNNNNN | 1 |
| Y | 6799 hCoV-19/Wales/PHWC-49AE49/2020\|EPI_ISL_727695\|2020 | ···················· | ···················· | ···················· | ···················· | NNNNNNNNNNNNNNNNNNNN | NNNNNNNNNNN·····NNNN | 1 |
| Y | 6800 hCoV-19/Wales/PHWC-49AE58/2020\|EPI_ISL_727696\|2020 | ···················· | ···················· | ···················· | ···················· | NNNNNNNNNNNNNNNNNNNN | NNNNNNNNNNNNNNNNNNNN | 1 |
| Y | 6801 hCoV-19/USA/WI-UW-2339/2020\|EPI_ISL_728105\|2020-11 | ···················· | ···················· | ···················· | ···················· | NNNNNNNNNNNNNNNNNNNN | NNNNNNNNNNNNNNNNNNNN | 1 |
| Y | 6802 hCoV-19/USA/WI-UW-2340/2020\|EPI_ISL_728106\|2020-11 | ···················· | ···················· | ···················· | ···················· | NNNNNNNNNNNNNNNNNNNN | NNNNNNNNNNNNNNNNNNNN | 1 |
| Y | 6803 hCoV-19/USA/IL-UW-2341/2020\|EPI_ISL_728107\|2020-11 | ···················· | ···················· | ···················· | ···················· | NNNNNNNNNNNNNNNNNNNN | NNNNNNNNNNNNNNNNNNNN | 1 |
| Y | 6805 hCoV-19/USA/WI-UW-2343/2020\|EPI_ISL_728109\|2020-11 | ···················· | ···················· | ···················· | ···················· | NNNNNNNNNNNNNNNNNNNN | NNNNNNNNNNNNNNNNNNNN | 1 |
| Y | 6920 hCoV-19/Wales/PHWC-4935A5/2020\|EPI_ISL_725909\|2020 | ···················· | ···················· | ···················· | ···················· | NNNNNNNNNNNNNNNNNNNN | ···················· | 1 |
| Y | 6921 hCoV-19/Wales/PHWC-4935B4/2020\|EPI_ISL_725910\|2020 | ···················· | ···················· | ···················· | ···················· | NNNNNNNNNNNNNNNNNNNN | NNNNNNNNNNNNNNNNNNNN | 1 |
| Y | 6923 hCoV-19/Wales/PHWC-4935E1/2020\|EPI_ISL_725913\|2020 | ···················· | ···················· | ···················· | ···················· | NNNNNNNNNNNNNNNNNNNN | NNNNNNNNNNNNNNNNNNNN | 1 |
| Y | 6927 hCoV-19/USA/WI-UW-2337/2020\|EPI_ISL_728103\|2020-11 | ···················· | ···················· | ···················· | ···················· | NNNNNNNNNNNNNNNNNNNN | NNNNNNNNNNNNNNNNNNNN | 1 |
| Y | 6928 hCoV-19/USA/WI-UW-2338/2020\|EPI_ISL_728104\|2020-11 | ···················· | ···················· | ···················· | ···················· | NNNNNNNNNNNNNNNNNNNN | NNNNNNNNNNNNNNNNNNNN | 1 |
| Y | 6932 hCoV-19/USA/WI-UW-2348/2020\|EPI_ISL_728114\|2020-11 | ···················· | ···················· | ···················· | ···················· | NNNNNNNNNNNNNNNNNNNN | NNNNNNNNNNNNNNNNNNNN | 1 |
| Y | 6935 hCoV-19/USA/WI-UW-2352/2020\|EPI_ISL_728118\|2020-11 | ···················· | ···················· | ···················· | ···················· | NNNNNNNNNNNNNNNNNNNN | NNNNNNNNNNNNNNNNNNNN | 1 |
| Y | 6936 hCoV-19/USA/WI-UW-2353/2020\|EPI_ISL_728119\|2020-11 | ···················· | ···················· | ···················· | ···················· | NNNNNNNNNNNNNNNNNNNN | NNNNNNNNNNNNNNNNNNNN | 1 |
| Y | 6938 hCoV-19/USA/WI-UW-2356/2020\|EPI_ISL_728122\|2020-11 | ···················· | ···T················ | ···················· | ···················· | NNNNNNNNNNNNNNNNNNNN | NNNNNNNNNNNNNNNNNNNN | 1 |
| Y | 6939 hCoV-19/USA/WI-UW-2358/2020\|EPI_ISL_728124\|2020-11 | ···················· | ···················· | ···················· | ···················· | NNNNNNNNNNNNNNNNNNNN | NNNNNNNNNNNNNNNNNNNN | 1 |
| Y | 6942 hCoV-19/USA/WI-UW-2361/2020\|EPI_ISL_728127\|2020-11 | ···················· | ···················· | ···················· | ···················· | NNNNNNNNNNNNNNNNNNNN | NNNNNNNNNNNNNNNNNNNN | 1 |
| Y | 6944 hCoV-19/USA/WI-UW-2363/2020\|EPI_ISL_728129\|2020-11 | ···················· | ···················· | ···················· | ···················· | NNNNNNNNNNNNNNNNNNNN | NNNNNNNNNNNNNNNNNNNN | 1 |
| Y | 6946 hCoV-19/USA/WI-UW-2365/2020\|EPI_ISL_728131\|2020-11 | ···················· | ···················· | ···················· | ···················· | NNNNNNNNNNNNNNNNNNNN | NNNNNNNNNNNNNNNNNNNN | 1 |
| Y | 6949 hCoV-19/USA/WI-UW-2368/2020\|EPI_ISL_728134\|2020-11 | ···················· | ···················· | ···················· | ···················· | NNNNNNNNNNNNNNNNNNNN | NNNNNNNNNNNNNNNNNNNN | 1 |
| Y | 6956 hCoV-19/USA/WI-UW-2375/2020\|EPI_ISL_728141\|2020-11 | ···················· | ···················· | ···················· | ···················· | NNNNNNNNNNNNNNNNNNNN | NNNNNNNNNNNNNNNNNNNN | 1 |

| | | | | | | | | |
|---|---|---|---|---|---|---|---|---|
| Y | 6957 hCoV-19/USA/WI-UW-2376/2020\|EPI_ISL_728142\|2020-10 | ···················· | ···················· | ···················· | ···················· | NNNNNNNNNNNNNNNNNNNN | NNNNNNNNNNNNNNNNNNNN | 1 |
| Y | 6958 hCoV-19/USA/WI-UW-2379/2020\|EPI_ISL_728144\|2020-11 | ···················· | ···················· | ···················· | ···················· | NNNNNNNNNNNNNNNNNNNN | NNNNNNNNNNNNNNNNNNNN | 1 |
| Y | 6989 hCoV-19/Germany/DE-SH-ChVir20046/2020\|EPI_ISL_7294 | ···················· | ···················· | ···················· | ···················· | ·······NNNNNNNNNNNNN | ···················· | 1 |
| Y | 7187 hCoV-19/Wales/PHWC-493684/2020\|EPI_ISL_725922\|2020 | ···················· | ···················· | ···················· | ···················· | ···············NNNNN | ···················· | 1 |
| Y | 7214 hCoV-19/Wales/PHWC-49388E/2020\|EPI_ISL_725951\|2020 | ···················· | ···················· | ···················· | ···················· | NNNNN··············· | ···················· | 1 |
| Y | 7219 hCoV-19/Wales/PHWC-4938F7/2020\|EPI_ISL_725956\|2020 | ···················· | ······A············· | ···················· | ···················· | NNNNNNNNNNNNNNNNNNNN | ···················· | 1 |
| Y | 7220 hCoV-19/Wales/PHWC-493903/2020\|EPI_ISL_725957\|2020 | ···················· | ···················· | ···················· | ···················· | NNNNNNNNNNNNNNNNN··· | NNNNNNNNNNNNNNNNNNNN | 1 |
| Y | 7223 hCoV-19/Wales/PHWC-49395E/2020\|EPI_ISL_725960\|2020 | ···················· | ···················· | ···················· | ···················· | NNNNNNNNNNNNNNNNNNNN | ···················· | 1 |
| Y | 7225 hCoV-19/Wales/PHWC-49398B/2020\|EPI_ISL_725963\|2020 | ···················· | ···················· | ···················· | ···················· | NNNNNNNNNNNNNNNNNNNN | ···················· | 1 |
| Y | 7233 hCoV-19/Wales/PHWC-493A2E/2020\|EPI_ISL_725973\|2020 | ···················· | ···················· | ···················· | ···················· | NNNN················ | ···················· | 1 |
| Y | 7238 hCoV-19/Wales/PHWC-493A79/2020\|EPI_ISL_725978\|2020 | ···················· | ···················· | ···················· | ···················· | NNNNNNNNNNNNNNNNNNNN | ···················· | 1 |
| Y | 7249 hCoV-19/Wales/PHWC-493BC1/2020\|EPI_ISL_725994\|2020 | ···················· | ···················· | ···················· | ···················· | ···················· | NNNNN··············· | 1 |
| Y | 7372 hCoV-19/Switzerland/ZH-ETHZ-410527/2020\|EPI_ISL_73 | ···················· | ···················· | ···················· | ···················· | ·······NNNNNNNNNNNNN | NNNNNNNNNNNNNNNNNNNN | 1 |
| Y | 7697 hCoV-19/Ecuador/USFQ-556/2020\|EPI_ISL_728204\|2020- | ···················· | ···················· | ···················· | ···················· | NNNNNNNNNNNNNNNNNNNN | NNNNNNNNN········NNN | 1 |
| Y | 7738 hCoV-19/Italy/LAZ-INMI-108/2020\|EPI_ISL_728281\|202 | ···················· | ···················· | ···················· | ···················· | NNNNNNNNNNNNNNN····· | ···················· | 1 |
| Y | 7740 hCoV-19/Italy/LAZ-INMI-110/2020\|EPI_ISL_728283\|202 | ···················· | ···················· | ···················· | ···················· | NNNNNNNNNNNNNNN····· | ···················· | 1 |
| Y | 7741 hCoV-19/Italy/LAZ-INMI-111/2020\|EPI_ISL_728284\|202 | ···················· | ···················· | ···················· | ···················· | NNNNNNNNNNNNNNN····· | ···················· | 1 |
| Y | 7862 hCoV-19/USA/MI-UM-10036820033/2020\|EPI_ISL_730566\| | ···················· | ·A·················· | ···················· | ···················· | NNNNNNNNNNNNNNNNNNNN | ···················· | 1 |
| Y | 7959 hCoV-19/France/ARA-105635/2020\|EPI_ISL_728304\|2020 | ···················· | ···················· | ···················· | ···················· | NNNNNNNNNNNNNNNNNNNN | ···················· | 1 |
| Y | 8141 hCoV-19/USA/WA-UW-31223/2020\|EPI_ISL_735464\|2020-1 | ···················· | ···················· | ···················· | ···················· | ···················· | ················NNNN | 1 |
| Y | 8199 hCoV-19/Wales/PHWC-497FB2/2020\|EPI_ISL_727007\|2020 | ···················· | ···················· | ···················· | ···················· | NNNNNNNNNNNNNNNNNNNN | ···················· | 1 |
| Y | 8202 hCoV-19/Wales/PHWC-497FEF/2020\|EPI_ISL_727010\|2020 | ···················· | ···················· | ···················· | ···················· | NNNNNNNNNNNNNNNNNNNN | NNNNNNNNNNNNNNNNNNNN | 1 |
| Y | 8218 hCoV-19/Wales/PHWC-498115/2020\|EPI_ISL_727028\|2020 | ···················· | ···················· | ···················· | ···················· | NNNNNNNNNNNNNNNNNNNN | NNNNNNNNNNNNNNNNNNNN | 1 |
| Y | 8275 hCoV-19/USA/MI-UM-10036804214/2020\|EPI_ISL_728374\| | ···················· | ····A··············· | ···················· | ···················· | NNNNNNNNNNNNNNNNNNNN | ···················· | 1 |
| Y | 8276 hCoV-19/USA/MI-UM-10036825013/2020\|EPI_ISL_728375\| | ···················· | ···················· | ···················· | ···················· | NNNNNNNNNNNNNNNNNNNN | ···················· | 1 |
| Y | 8291 hCoV-19/Germany/DE-BE-ChVir8991/2020\|EPI_ISL_72960 | ···················· | ···················· | ···················· | ···················· | NNNNNNNNNNNNNNNNNNNN | ···················· | 1 |
| Y | 8321 hCoV-19/Germany/DE-SH-ChVir9989/2020\|EPI_ISL_72966 | ···················· | ···················· | ···················· | ···················· | ······NNNNNNNNNNNNNN | ···················· | 1 |
| Y | 8322 hCoV-19/Germany/DE-SH-ChVir20035/2020\|EPI_ISL_7296 | ···················· | ···················· | ···················· | ···················· | ······NNNNNNNNNNNNNN | ···················· | 1 |
| Y | 8492 hCoV-19/Wales/PHWC-498151/2020\|EPI_ISL_727032\|2020 | ···················· | ···················· | ···················· | ···················· | NNNNNNNNNNNNNNNNNNNN | NNNNNNNNNNNNNNNNNNNN | 1 |
| Y | 8493 hCoV-19/Wales/PHWC-498160/2020\|EPI_ISL_727033\|2020 | ···················· | ···················· | ···················· | ···················· | NNNNNNNNNNNNNNNNNNNN | ···················· | 1 |
| Y | 8531 hCoV-19/Wales/PHWC-4985CE/2020\|EPI_ISL_727097\|2020 | ···················· | ···················· | ···················· | ···················· | ···················· | NNNNNNNNNNNNNNN····· | 1 |
| Y | 8533 hCoV-19/Wales/PHWC-4985EC/2020\|EPI_ISL_727099\|2020 | ···················· | ···················· | ···················· | ···················· | NNNNNNNNNNNNNNNNNNNN | NNNNNNNNNNNNNNNNNNNN | 1 |
| Y | 8543 hCoV-19/USA/MI-UM-10036816999/2020\|EPI_ISL_728391\| | ···················· | ···················· | ···················· | ···················· | NNNNNNNNNNNNNNNNNNNN | ···················· | 1 |
| Y | 8712 hCoV-19/Wales/PHWC-4985FB/2020\|EPI_ISL_727100\|2020 | ···················· | ···················· | ···················· | ···················· | NNNNNNNNNNNNNNNNNNNN | NNNNNNNNNNNNNNNNNNNN | 1 |
| Y | 8715 hCoV-19/Wales/PHWC-498643/2020\|EPI_ISL_727104\|2020 | ···················· | ···················· | ···················· | ···················· | NNNNNNNNNNNNNNNNNNNN | NNNNNNNNNNNNNNNNNNNN | 1 |
| Y | 8716 hCoV-19/Wales/PHWC-498652/2020\|EPI_ISL_727105\|2020 | ···················· | ···················· | ···················· | ···················· | ···················· | NNNNNNNNNNNNNNN····· | 1 |
| Y | 8717 hCoV-19/Wales/PHWC-498661/2020\|EPI_ISL_727106\|2020 | ···················· | ···················· | ···················· | ···················· | NNNNNNNNNNNNNNNNNNNN | NNNNNNNNNNNNNNNNNNNN | 1 |
| Y | 8720 hCoV-19/Wales/PHWC-49869E/2020\|EPI_ISL_727109\|2020 | ···················· | ···················· | ···················· | ···················· | NNNNNNNNNNNNNNNNNNNN | ···················· | 1 |
| Y | 8723 hCoV-19/Wales/PHWC-4987D7/2020\|EPI_ISL_727124\|2020 | ···················· | ···················· | ···················· | ···················· | NNNNNNNNNNNNNNNNNNNN | ···················· | 1 |
| Y | 8726 hCoV-19/Wales/PHWC-49887A/2020\|EPI_ISL_727133\|2020 | ···················· | ···················· | ···················· | ···················· | NNNNNNNNNNNNNNNNNNNN | ···················· | 1 |
| Y | 8727 hCoV-19/Wales/PHWC-4988A7/2020\|EPI_ISL_727136\|2020 | ···················· | ···················· | ···················· | ···················· | ···················· | ·····NNNNNNNNNNNNNNN | 1 |
| Y | 8729 hCoV-19/Wales/PHWC-4988D4/2020\|EPI_ISL_727139\|2020 | ···················· | ···················· | ···················· | ···················· | NNNNNNNNNNNNNNNNNNNN | ···················· | 1 |
| Y | 8737 hCoV-19/USA/MI-UM-10036820004/2020\|EPI_ISL_728409\| | ···················· | ···················· | ···················· | ···················· | NNNNNNNNNNNNNNNNNNNN | ···················· | 1 |
| Y | 8739 hCoV-19/USA/MI-UM-10036813449/2020\|EPI_ISL_728411\| | ···················· | ·············A······ | ···················· | ···················· | NNNNNNNNNNNNNNNNNNNN | NNNNNNNNNNNNNNNNNNNN | 1 |
| Y | 8740 hCoV-19/USA/MI-UM-10036816239/2020\|EPI_ISL_728412\| | ···················· | ···················· | ···················· | ···················· | NNNNNNNNNNNNNNNNNNNN | ···················· | 1 |
| Y | 8741 hCoV-19/USA/MI-UM-10036817758/2020\|EPI_ISL_728413\| | ···················· | ···················· | ···················· | ···················· | NNNNNNNNNNNNNNNNNNNN | ···················· | 1 |
| Y | 8744 hCoV-19/USA/MI-UM-10036814157/2020\|EPI_ISL_728416\| | ···················· | ···················· | ···················· | ···················· | NNNNNNNNNNNNNNNNNNNN | ···················· | 1 |
| Y | 8745 hCoV-19/USA/MI-UM-10036820127/2020\|EPI_ISL_728417\| | ···················· | ···················· | ···················· | ···················· | NNNNNNNNNNNNNNNNNNNN | ···················· | 1 |
| Y | 8746 hCoV-19/USA/MI-UM-10036824672/2020\|EPI_ISL_728418\| | ···················· | ···················· | ···················· | ···················· | NNNNNNNNNNNNNNNNNNNN | NNNNNNNNNNNNNNNNNNNN | 1 |
| Y | 8747 hCoV-19/USA/MI-UM-10036825929/2020\|EPI_ISL_728419\| | ···················· | ···················· | ···················· | ···················· | NNNNNNNNNNNNNNNNNNNN | NNNNNNNNNNNNNNNNNNNN | 1 |
| Y | 8748 hCoV-19/USA/MI-UM-10036816620/2020\|EPI_ISL_728421\| | ···················· | ···················· | ···················· | ···················· | NNNNNNNNNNNNNNNNNNNN | ···················· | 1 |
| Y | 8749 hCoV-19/USA/MI-UM-10036817603/2020\|EPI_ISL_728422\| | ···················· | ···················· | ···················· | ···················· | NNNNNNNNNNNNNNNNNNNN | NNNNNNNNNNNNNNNNNNNN | 1 |
| Y | 8751 hCoV-19/USA/MI-UM-10036825916/2020\|EPI_ISL_728426\| | ···················· | ·············A······ | ···················· | ···················· | NNNNNNNNNNNNNNNNNNNN | ···················· | 1 |
| Y | 8756 hCoV-19/USA/MI-UM-10036825275/2020\|EPI_ISL_728431\| | ···················· | ···················· | ···················· | ···················· | NNNNNNNNNNNNNNNNNNNN | ···················· | 1 |
| Y | 8760 hCoV-19/USA/MI-UM-10036835049/2020\|EPI_ISL_728437\| | ···················· | ···················· | ···················· | ···················· | NNNNNNNNNNNNNNNNNNNN | ···················· | 1 |
| Y | 8762 hCoV-19/USA/MI-UM-10036834956/2020\|EPI_ISL_728439\| | ···················· | ···················· | ···················· | ···················· | NNNNNNNNNNNNNNNNNNNN | NNNNNNNNNNNNNNNNNNNN | 1 |
| Y | 8763 hCoV-19/USA/MI-UM-10036850459/2020\|EPI_ISL_728450\| | ···················· | ···················· | ···················· | ···················· | NNNNNNNNNNNNNNNNNNNN | NNNNNNNNNNNNNNNNNNNN | 1 |
| Y | 8764 hCoV-19/USA/MI-UM-10036847408/2020\|EPI_ISL_728451\| | ···················· | ···················· | ···················· | ···················· | NNNNNNNNNNNNNNNNNNNN | ···················· | 1 |
| Y | 8765 hCoV-19/USA/MI-UM-10036855102/2020\|EPI_ISL_728452\| | ···················· | ···················· | ···················· | ···················· | NNNNNNNNNNNNNNNNNNNN | ···················· | 1 |
| Y | 8766 hCoV-19/USA/MI-UM-10036858527/2020\|EPI_ISL_728453\| | ···················· | ···················· | ···················· | ···················· | NNNNNNNNNNNNNNNNNNNN | ···················· | 1 |
| Y | 8767 hCoV-19/USA/MI-UM-10036857041/2020\|EPI_ISL_728454\| | ···················· | ···················· | ···················· | ···················· | NNNNNNNNNNNNNNNNNNNN | ···················· | 1 |
| Y | 8769 hCoV-19/USA/MI-UM-10036858504/2020\|EPI_ISL_728456\| | ···················· | ···················· | ···················· | ···················· | NNNNNNNNNNNNNNNNNNNN | NNNNNNNNNNNNNNNNNNNN | 1 |
| Y | 8770 hCoV-19/USA/MI-UM-10036836329/2020\|EPI_ISL_728457\| | ···················· | ···················· | ···················· | ···················· | NNNNNNNNNNNNNNNNNNNN | ···················· | 1 |
| Y | 8771 hCoV-19/USA/MI-UM-10036836424/2020\|EPI_ISL_728460\| | ···················· | ·A·················· | ···················· | ···················· | NNNNNNNNNNNNNNNNNNNN | ···················· | 1 |
| Y | 8772 hCoV-19/USA/MI-UM-10036835773/2020\|EPI_ISL_728461\| | ···················· | ···················· | ···················· | ···················· | NNNNNNNNNNNNNNNNNNNN | NNNNNNNNNNNNNNNNNNNN | 1 |
| Y | 8777 hCoV-19/USA/MI-UM-10036835872/2020\|EPI_ISL_728468\| | ···················· | ···················· | ···················· | ···················· | NNNNNNNNNNNNNNNNNNNN | NNNNNNNNNNNNNNNNNNNN | 1 |
| Y | 8778 hCoV-19/USA/MI-UM-10036832946/2020\|EPI_ISL_728469\| | ···················· | ·············A······ | ···················· | ···················· | NNNNNNNNNNNNNNNNNNNN | ···················· | 1 |
| Y | 8985 hCoV-19/Wales/PHWC-4986BC/2020\|EPI_ISL_727111\|2020 | ···················· | ···················· | ···················· | ···················· | NNNNNNNNNNNNNNNNNNNN | NNNNNNNNNNNNNNNNNNNN | 1 |
| Y | 8986 hCoV-19/Wales/PHWC-4986CB/2020\|EPI_ISL_727112\|2020 | ···················· | ···················· | ···················· | ···················· | ···················· | NNNNNNNN············ | 1 |
| Y | 8988 hCoV-19/Wales/PHWC-498713/2020\|EPI_ISL_727116\|2020 | ···················· | ···················· | ···················· | ···················· | NNNNNNNNNNNNNNNNNNNN | NNNNNNNNNNNNNNNNNNNN | 1 |
| Y | 8989 hCoV-19/Wales/PHWC-498722/2020\|EPI_ISL_727117\|2020 | ···················· | ···················· | ···················· | ···················· | NNNNNNNNNNNNNNNNNNNN | NNNNNNNNNNNNNNNNNNNN | 1 |
| Y | 8992 hCoV-19/Wales/PHWC-4988E3/2020\|EPI_ISL_727140\|2020 | ···················· | ···················· | ···················· | ···················· | NNNNNNNNNNNNNNNNNNNN | NNNNNNNNNNNNNNNNNNNN | 1 |
| Y | 8995 hCoV-19/Wales/PHWC-49891D/2020\|EPI_ISL_727143\|2020 | ···················· | ···················· | ···················· | ···················· | ···················· | NNNNN··············· | 1 |
| Y | 8996 hCoV-19/Wales/PHWC-49892C/2020\|EPI_ISL_727144\|2020 | ···················· | ···················· | ···················· | ···················· | ··NNNNNNNNNNNNNNN··· | ····N······NNNN····· | 1 |
| Y | 8998 hCoV-19/Wales/PHWC-49894A/2020\|EPI_ISL_727146\|2020 | ···················· | ···················· | ···················· | ···················· | NNNNNNNNNNNNNNNNNNNN | NNNNNNNNNNNNNNNNNNNN | 1 |
| Y | 8999 hCoV-19/Wales/PHWC-498968/2020\|EPI_ISL_727148\|2020 | ···················· | ···················· | ···················· | ···················· | NNNNNNNNNNNNNNNNNNNN | NNNNNNNNNNNNNNNNNNNN | 1 |
| Y | 9000 hCoV-19/Wales/PHWC-498977/2020\|EPI_ISL_727149\|2020 | ···················· | ···················· | ···················· | ···················· | NNNNNNNNNNNNNNNNNNNN | NNNNNNNNNNNNNNNNNNNN | 1 |
| Y | 9001 hCoV-19/Wales/PHWC-498986/2020\|EPI_ISL_727150\|2020 | ···················· | ···················· | ···················· | ···················· | ···················· | NNNNNNNNNNNNNNN····· | 1 |
| Y | 9003 hCoV-19/Wales/PHWC-4989A4/2020\|EPI_ISL_727152\|2020 | ···················· | ···················· | ···················· | ···················· | NNNNNNNNNNNNNNNNNNNN | NNNNNNNNNNNNNNNNNNNN | 1 |
| Y | 9007 hCoV-19/Wales/PHWC-498A0B/2020\|EPI_ISL_727158\|2020 | ···················· | ···················· | ···················· | ···················· | NNNNNNNNNNNNNNNNNNNN | NNNNNNNNNNNNNNNNNNNN | 1 |

| | | | | | | | | | |
|---|---|---|---|---|---|---|---|---|---|
| Y | 9013 | hCoV-19/Wales/PHWC-498A74/2020\|EPI_ISL_727165\|2020 | ···················· | ···················· | ···················· | ···················· | NNNNNNNNNNNNNNNNNNNN | NNNNNNNNNNNNNNNNNNNN | 1 |
| Y | 9015 | hCoV-19/Wales/PHWC-498AA1/2020\|EPI_ISL_727168\|2020 | ···················· | ···················· | ···················· | ···················· | NNNNNNNNNNNNNNNNNNNN | NNNNNNNNNNNNNNNNNNNN | 1 |
| Y | 9016 | hCoV-19/Wales/PHWC-498ADE/2020\|EPI_ISL_727171\|2020 | ···················· | ···················· | ···················· | ···················· | NNNNNNNNNNNNNNNNNNNN | NNNNNNNNNNNNNNNNNNNN | 1 |
| Y | 9017 | hCoV-19/Wales/PHWC-498AED/2020\|EPI_ISL_727172\|2020 | ···················· | ···················· | ···················· | ···················· | NNNNNNNNNNNNNNNNNNNN | NNNNNNNNNNNNNNN··NNN | 1 |
| Y | 9019 | hCoV-19/Wales/PHWC-498B53/2020\|EPI_ISL_727178\|2020 | ···················· | ···················· | ···················· | ···················· | NNNNNNNNNNNNNNNNNNNN | NNNNNNNNNNNNNNNNNNNN | 1 |
| Y | 9020 | hCoV-19/Wales/PHWC-498B62/2020\|EPI_ISL_727179\|2020 | ···················· | ···················· | ···················· | ···················· | NNNNNNNNNNNNNNNNNNNN | NNNNNNNNNNNNNNNNNNNN | 1 |
| Y | 9025 | hCoV-19/Wales/PHWC-498C23/2020\|EPI_ISL_727189\|2020 | ···················· | ···················· | ···················· | ···················· | NNNNNNNNNNNNNNNNNNNN | NNNNNNNNNNNNNNNNNNNN | 1 |
| Y | 9026 | hCoV-19/Wales/PHWC-498C32/2020\|EPI_ISL_727190\|2020 | ···················· | ···················· | ···················· | ···················· | NNNNNNNNNNNNNNNNNNNN | ·········NNNNNNNNNNN | 1 |
| Y | 9028 | hCoV-19/Wales/PHWC-498C50/2020\|EPI_ISL_727192\|2020 | ···················· | ···················· | ···················· | ···················· | NNNNNNNNNNNNNNNNNNNN | NNNNNNNNNNNNNNNNNNNN | 1 |
| Y | 9029 | hCoV-19/Wales/PHWC-498C6F/2020\|EPI_ISL_727193\|2020 | ···················· | ···················· | ···················· | ···················· | NNNNNNNNNNNNNNNNNNNN | ···················· | 1 |
| Y | 9030 | hCoV-19/Wales/PHWC-498C8D/2020\|EPI_ISL_727194\|2020 | ···················· | ···················· | ···················· | ···················· | NNNNNNNNNNNN········ | ···················· | 1 |
| Y | 9031 | hCoV-19/Wales/PHWC-498CBA/2020\|EPI_ISL_727197\|2020 | ···················· | ···················· | ···················· | ···················· | NNNNNNNNNNNNNNNNNNNN | NNNNNNNNNNNNNNNNNNNN | 1 |
| Y | 9032 | hCoV-19/USA/MI-UM-10036833206/2020\|EPI_ISL_728440\| | ···················· | ···················· | ···················· | ···················· | NNNNNNNNNNNNNNNNNNNN | ···················· | 1 |
| Y | 9035 | hCoV-19/USA/MI-UM-10036834888/2020\|EPI_ISL_728443\| | ···················· | ···················· | ···················· | ···················· | NNNNNNNNNNNNNNNNNNNN | NNNNNNNNNNNNNNNNNNNN | 1 |
| Y | 9038 | hCoV-19/USA/MI-UM-10036852325/2020\|EPI_ISL_728446\| | ···················· | ···················· | ···················· | ···················· | NNNNNNNNNNNNNNNNNNNN | NNNNNNNNNNNNNNNNNNNN | 1 |
| Y | 9040 | hCoV-19/USA/MI-UM-10036855179/2020\|EPI_ISL_728448\| | ···················· | ···················· | ···················· | ···················· | NNNNNNNNNNNNNNNNNNNN | NNNNNNNNNNNNNNNNNNNN | 1 |
| Y | 9044 | hCoV-19/USA/MI-UM-10036852759/2020\|EPI_ISL_728476\| | ···················· | ·············A······ | ···················· | ···················· | NNNNNNNNNNNNNNNNNNNN | NNNNNNNNNNNNNNNNNNNN | 1 |
| Y | 9047 | hCoV-19/USA/MI-UM-10036873336/2020\|EPI_ISL_728482\| | ···················· | ···················· | ···················· | ···················· | NNNNNNNNNNNNNNNNNNNN | NNNNNNNNNNNNNNNNNNNN | 1 |
| Y | 9048 | hCoV-19/USA/MI-UM-10036858413/2020\|EPI_ISL_728483\| | ···················· | ·A·················· | ···················· | ···················· | NNNNNNNNNNNNNNNNNNNN | ···················· | 1 |
| Y | 9049 | hCoV-19/USA/MI-UM-10036876596/2020\|EPI_ISL_728485\| | ···················· | ···················· | ···················· | ···················· | NNNNNNNNNNNNNNNNNNNN | NNNNNNNNNNNNNNNNNNNN | 1 |
| Y | 9050 | hCoV-19/USA/MI-UM-10036871836/2020\|EPI_ISL_728486\| | ···················· | ···················· | ···················· | ···················· | NNNNNNNNNNNNNNNNNNNN | NNNNNNNNNNNNNNNNNNNN | 1 |
| Y | 9053 | hCoV-19/USA/MI-UM-10036857965/2020\|EPI_ISL_728490\| | ···················· | ···················· | ···················· | ···················· | NNNNNNNNNNNNNNNNNNNN | ···················· | 1 |
| Y | 9054 | hCoV-19/USA/MI-UM-10036849861/2020\|EPI_ISL_728491\| | ···················· | ···················· | ···················· | ···················· | NNNNNNNNNNNNNNNNNNNN | NNNNNNNNNNNNNNNNNNNN | 1 |
| Y | 9056 | hCoV-19/USA/MI-UM-10036853496/2020\|EPI_ISL_728494\| | ···················· | ···················· | ···················· | ···················· | NNNNNNNNNNNNNNNNNNNN | NNNNNNNNNNNNNNNNNNNN | 1 |
| Y | 9057 | hCoV-19/USA/MI-UM-10036852618/2020\|EPI_ISL_728495\| | ···················· | ···················· | ···················· | ···················· | NNNNNNNNNNNNNNNNNNNN | ···················· | 1 |
| Y | 9059 | hCoV-19/USA/MI-UM-10036853506/2020\|EPI_ISL_728497\| | ···················· | ···················· | ···················· | ···················· | ··NNNNNNNNNNNNNNNNNN | ···················· | 1 |
| Y | 9060 | hCoV-19/USA/MI-UM-10036849862/2020\|EPI_ISL_728499\| | ···················· | ···················· | ···················· | ···················· | NNNNNNNNNNNNNNNNNNNN | NNNNNNNNNNNNNNNNNNNN | 1 |
| Y | 9202 | hCoV-19/South Africa/KRISP-WC-SA04352428/2020\|EPI_ | ···················· | ···················· | ···················· | ···················· | ··NNNNNNNNNNNNNNNNNN | NNNNNNNNNNNNNNNNNNNN | 1 |
| Y | 9218 | hCoV-19/Wales/PHWC-498D3F/2020\|EPI_ISL_727203\|2020 | ···················· | ···················· | ···················· | ···················· | NNNNNNNNNNNNNNNNNNNN | NNNNNNNNNNNNNNNNNNNN | 1 |
| Y | 9219 | hCoV-19/Wales/PHWC-498D4E/2020\|EPI_ISL_727204\|2020 | ···················· | ···················· | ···················· | ···················· | NNNNNNNNNNNNNNNNNNNN | NNNNNNNNNNNNNNN·NNNN | 1 |
| Y | 9221 | hCoV-19/Wales/PHWC-498D8A/2020\|EPI_ISL_727207\|2020 | ···················· | ···················· | ···················· | ···················· | NNNNNNNNNNNNNNNNNNNN | NNNNNNNNNNNNNNNNNNNN | 1 |
| Y | 9222 | hCoV-19/Wales/PHWC-498DA8/2020\|EPI_ISL_727209\|2020 | ···················· | ···················· | ···················· | ···················· | NNNNNNNNNNNNNNNNNNNN | NNNNNNNNNNNNNNNNNNNN | 1 |
| Y | 9223 | hCoV-19/Wales/PHWC-498DB7/2020\|EPI_ISL_727210\|2020 | ···················· | ···················· | ···················· | ···················· | NNNNNNNNNNNNNNNNNNNN | NNNNNNNNNNNNNNNNNNNN | 1 |
| Y | 9225 | hCoV-19/Wales/PHWC-498DD5/2020\|EPI_ISL_727212\|2020 | ···················· | ···················· | ···················· | ···················· | NNNNNNNNNNNNNNNNN··· | ···················· | 1 |
| Y | 9228 | hCoV-19/Wales/PHWC-498E1E/2020\|EPI_ISL_727215\|2020 | ···················· | ···················· | ···················· | ···················· | NNNNNNNNNNNNNNNNNNNN | NNNNN······NNNNNNNNN | 1 |
| Y | 9229 | hCoV-19/Wales/PHWC-498E3C/2020\|EPI_ISL_727217\|2020 | ···················· | ···················· | ···················· | ···················· | NNNNNNNNNNNNNNNNNNNN | NNNNNNNNNNNNNNNNNNNN | 1 |
| Y | 9231 | hCoV-19/Wales/PHWC-498E5A/2020\|EPI_ISL_727219\|2020 | ···················· | ···················· | ···················· | ···················· | NNNNNNNNNNNNNNNNNNNN | NNNNNNNNNNNNNNNNNNNN | 1 |
| Y | 9232 | hCoV-19/USA/MI-UM-10036858924/2020\|EPI_ISL_728500\| | ···················· | ···················· | ···················· | ···················· | NNNNNNNNNNNNNNNNNNNN | ···················· | 1 |
| Y | 9234 | hCoV-19/USA/MI-UM-10036855767/2020\|EPI_ISL_728502\| | ···················· | ···················· | ···················· | ···················· | NNNNNNNNNNNNNNNNNNNN | ···················· | 1 |
| Y | 9235 | hCoV-19/USA/MI-UM-10036852112/2020\|EPI_ISL_728504\| | ···················· | ···················· | ···················· | ···················· | NNNNNNNNNNNNNNNNNNNN | NNNNNNNNNNNNNNNNNNNN | 1 |
| Y | 9236 | hCoV-19/USA/MI-UM-10036849897/2020\|EPI_ISL_728505\| | ···················· | ···················· | ···················· | ···················· | NNNNNNNNNNNNNNNNNNNN | NNNNNNNNNNNNNNNNNNNN | 1 |
| Y | 9237 | hCoV-19/USA/MI-UM-10036861068/2020\|EPI_ISL_728506\| | ···················· | ·A·················· | ···················· | ···················· | NNNNNNNNNNNNNNNNNNNN | ···················· | 1 |
| Y | 9238 | hCoV-19/USA/MI-UM-10036855279/2020\|EPI_ISL_728507\| | ···················· | ···················· | ···················· | ···················· | NNNNNNNNNNNNNNNNNNNN | ···················· | 1 |
| Y | 9239 | hCoV-19/USA/MI-UM-10036852629/2020\|EPI_ISL_728508\| | ···················· | ···················· | ···················· | ···················· | ······NNNNNNNNNNNNNN | ···················· | 1 |
| Y | 9240 | hCoV-19/USA/MI-UM-10036876426/2020\|EPI_ISL_728509\| | ···················· | ···················· | ···················· | ···················· | NNNNNNNNNNNNNNNNNNNN | ···················· | 1 |
| Y | 9242 | hCoV-19/USA/MI-UM-10036861408/2020\|EPI_ISL_728513\| | ···················· | ···················· | ···················· | ···················· | NNNNNNNNNNNNNNNNNNNN | ···················· | 1 |
| Y | 9243 | hCoV-19/USA/MI-UM-10036859255/2020\|EPI_ISL_728515\| | ···················· | ···················· | ···················· | ···················· | NNNNNNNNNNNNNNNNNNNN | NNNNNNNNNNNNNNNNNNNN | 1 |
| Y | 9423 | hCoV-19/Russia/SVE-RII-MH3110S/2020\|EPI_ISL_733089 | ···················· | ···················· | ···················· | ···················· | ···················· | ····NNNNNNNNNNNNNNNN | 1 |
| Z | 21 | hCoV-19/Wales/PHWC-49FAAA/2020\|EPI_ISL_743315\|2020 | ···················· | ···················· | ···················· | ···················· | NNNNNNNNNNNNNNNNNNNN | NNNNNNNNNNNNNNNNNNNN | 1 |
| Z | 32 | hCoV-19/Wales/PHWC-49FBB6/2020\|EPI_ISL_743328\|2020 | ···················· | ···················· | ···················· | ···················· | NNNNNNNNNNNNN······· | ···················· | 1 |
| Z | 45 | hCoV-19/Wales/PHWC-49FCE0/2020\|EPI_ISL_743344\|2020 | ···················· | ···················· | ···················· | ···················· | NNNNNNNNNNNNNNNNNNNN | NNNNNNNNNN·········· | 1 |
| Z | 48 | hCoV-19/Wales/PHWC-49FD1A/2020\|EPI_ISL_743347\|2020 | ···················· | ···················· | ···················· | ···················· | NNNNNNNNNNNNNNNNNNNN | NNNNNNNNNNNNNNNNNNNN | 1 |
| Z | 50 | hCoV-19/Wales/PHWC-49FD47/2020\|EPI_ISL_743349\|2020 | ···················· | ···················· | ···················· | ···················· | ···················· | NNNNNNNNN··········· | 1 |
| Z | 52 | hCoV-19/Wales/PHWC-49FDB0/2020\|EPI_ISL_743355\|2020 | ···················· | ···················· | ···················· | ···················· | NNNNNNNNNNNNNNNNNNNN | ···················· | 1 |
| Z | 54 | hCoV-19/Wales/PHWC-49FE35/2020\|EPI_ISL_743359\|2020 | ···················· | ···················· | ···················· | ···················· | NNNNNNNNNNNNNNNNNNNN | NNNNNNNNNNNNNNNNNNNN | 1 |
| Z | 55 | hCoV-19/Wales/PHWC-49FE71/2020\|EPI_ISL_743362\|2020 | ···················· | ···················· | ···················· | ···················· | NNNNNNNNNNNNNNNNNNNN | NNNNNNNNNNNNNNNNNNNN | 1 |
| Z | 59 | hCoV-19/Wales/PHWC-49FF05/2020\|EPI_ISL_743369\|2020 | ···················· | ···················· | ···················· | ···················· | NNNNNNNNNNNNNNNNNNNN | NNNNNNNNNNNNNNNNNNNN | 1 |
| Z | 60 | hCoV-19/Wales/PHWC-49FF14/2020\|EPI_ISL_743370\|2020 | ···················· | ···················· | ···················· | ···················· | ······NNNNNNNNNNNNNN | ···················· | 1 |
| Z | 61 | hCoV-19/Wales/PHWC-49FF23/2020\|EPI_ISL_743371\|2020 | ···················· | ···················· | ···················· | ···················· | NNNNNNNNNNNNNNNNNNNN | ···················· | 1 |
| Z | 259 | hCoV-19/USA/CA-CZB-14430/2020\|EPI_ISL_738802\|2020- | ···················· | ···················· | ···················· | ···················· | NNNNNNNNNNNNNNNNNNNN | NNNNNNNNNNNNNNNNNNNN | 1 |
| Z | 269 | hCoV-19/USA/CA-CZB-15167/2020\|EPI_ISL_738812\|2020- | ···················· | ·A·················· | ···················· | ···················· | NNNNNNNNNNNNNNNNNNNN | ···················· | 1 |
| Z | 270 | hCoV-19/USA/CA-CZB-14322/2020\|EPI_ISL_738813\|2020- | ···················· | ···················· | ·········T·········· | ···················· | NNNNNNNNNNNNNNNNNNNN | ···················· | 1 |
| Z | 271 | hCoV-19/USA/CA-CZB-13790/2020\|EPI_ISL_738814\|2020- | ···················· | ·A·················· | ···················· | ···················· | NNNNNNNNNNNNNNNNNNNN | NNNNNNNNNNNNNNNNNNNN | 1 |
| Z | 276 | hCoV-19/USA/CA-CZB-14323/2020\|EPI_ISL_738819\|2020- | ···················· | ···················· | ···················· | ···················· | NNNNNNNNNNNNNN······ | ···················· | 1 |
| Z | 288 | hCoV-19/USA/CA-CZB-13829/2020\|EPI_ISL_738831\|2020- | ···················· | ·A·················· | ···················· | ···················· | ···················· | NNNNNNNNNNNNNNN····· | 1 |
| Z | 290 | hCoV-19/USA/CA-CZB-13813/2020\|EPI_ISL_738833\|2020- | ···················· | ···················· | ···················· | ···················· | NNNNNNNNNNNNNNNNNNNN | NNNNNNNNNNNNNNNNNNNN | 1 |
| Z | 296 | hCoV-19/USA/CA-CZB-15266/2020\|EPI_ISL_738839\|2020- | ···················· | ···················· | ···················· | ···················· | NNNNNNNNNNNNNNNNNNNN | NNNNNNNNNNNNNNN····· | 1 |
| Z | 303 | hCoV-19/USA/CA-CZB-14262/2020\|EPI_ISL_738846\|2020- | ···················· | ···················· | ···················· | ···················· | NNNNNNNNNNNNNNNNNNNN | NNNNNNNNNNNNNNNNNNNN | 1 |
| Z | 318 | hCoV-19/USA/CA-CZB-14563/2020\|EPI_ISL_738861\|2020- | ···················· | ···················· | ···················· | ···················· | ·NN······NNNNNNNNNNN | NNNNNNNNNNNNNNNNNNNN | 1 |
| Z | 340 | hCoV-19/USA/CA-CZB-15302/2020\|EPI_ISL_738883\|2020- | ···················· | ·A·················· | ···················· | ···················· | NNNNNNNNNNNNNNNNNNNN | NNNNNNNNNNNNNNNNNNNN | 1 |
| Z | 343 | hCoV-19/USA/CA-CZB-14097/2020\|EPI_ISL_738886\|2020- | ···················· | ···················· | ···················· | ···················· | ··NNNNNNNNNNNN······ | R··················· | 1 |
| Z | 344 | hCoV-19/USA/CA-CZB-14582/2020\|EPI_ISL_738887\|2020- | ···················· | ···················· | ···················· | ···················· | NNNNNNNNNNNNNNNNNNNN | ···················· | 1 |
| Z | 355 | hCoV-19/USA/CA-CZB-14567/2020\|EPI_ISL_738898\|2020- | ···················· | ···················· | ···················· | ···················· | ···················· | NNNNNNNNN··········· | 1 |
| Z | 382 | hCoV-19/Wales/PHWC-49FFD8/2020\|EPI_ISL_743381\|2020 | ···················· | ···················· | ···················· | ···················· | NNNNNNNNNNNNNNNNNNNN | NNNNNNNNNNNNNNNNNNNN | 1 |
| Z | 383 | hCoV-19/Wales/PHWC-4A002C/2020\|EPI_ISL_743385\|2020 | ···················· | ···················· | ···················· | ···················· | NNNNNNNNNNNNNNNNNNNN | NNNNNNNNNNNNNNNNNNNN | 1 |
| Z | 384 | hCoV-19/Wales/PHWC-4A003B/2020\|EPI_ISL_743386\|2020 | ···················· | ···················· | ···················· | ···················· | NNNNNNNNNNNNNNNNNNNN | NNNNN·NN·········NNN | 1 |
| Z | 385 | hCoV-19/Wales/PHWC-4A004A/2020\|EPI_ISL_743387\|2020 | ···················· | ···················· | ···················· | ···················· | NNNNNNNNNNNNNNNNNNNN | NNNNNNNNNNNNNNNNNNNN | 1 |
| Z | 386 | hCoV-19/Wales/PHWC-4A0068/2020\|EPI_ISL_743389\|2020 | ···················· | ···················· | ···················· | ···················· | NNNNNNNNNNNNNNNNNNNN | NNNNNNNNNNNNNNNNNNNN | 1 |
| Z | 389 | hCoV-19/Wales/PHWC-4A00C2/2020\|EPI_ISL_743394\|2020 | ···················· | ···················· | ···················· | ···················· | NNNNNNNNNNNNNNNNNNNN | ···················· | 1 |
| Z | 390 | hCoV-19/Wales/PHWC-4A00D1/2020\|EPI_ISL_743395\|2020 | ···················· | ···················· | ···················· | ···················· | NNNNNNNNNNNNNNNNNNNN | NNNNNNNNNNNNNNNNNNNN | 1 |

```
Z 391 hCoV-19/Wales/PHWC-4A00E0/2020|EPI_ISL_743396|2020 ···················· ···················· ···················· ···················· NNNNNNNNNNNNNNNNNNNN NNNNNNNNNNNNNNNNNNNN 1
Z 393 hCoV-19/Wales/PHWC-4A0138/2020|EPI_ISL_743400|2020 ···················· ···················· ···················· ···················· NNNNNNNNNNNNNNNNNNNN NNNNNNNNNNNNNNNNNNNN 1
Z 413 hCoV-19/Wales/PHWC-4A036F/2020|EPI_ISL_743434|2020 ···················· ···················· ···················· ···················· NNNNNNNNNNNNNNNN···· NNNNNNNNNNNNNNNNNNNN 1
Z 415 hCoV-19/Wales/PHWC-4A039C/2020|EPI_ISL_743437|2020 ···················· ···················· ···················· ···················· NNNNN···NNNNNNNNNNNN ···················· 1
Z 416 hCoV-19/Wales/PHWC-4A03BA/2020|EPI_ISL_743439|2020 ···················· ···················· ···················· ···················· NNNNNNNNNNNNNNNNNNNN NNNNNNNNNNNNNNNNNNNN 1
Z 417 hCoV-19/Wales/PHWC-4A03E7/2020|EPI_ISL_743440|2020 ···················· ···················· ···················· ···················· NNNNNNNNNNNNNNNNNNNN NNNNNNNNNNNNNNNNNNNN 1
Z 424 hCoV-19/Wales/PHWC-4A04C6/2020|EPI_ISL_743453|2020 ···················· ···················· ···················· ···················· NNNNNNNNNNNN········ NN·················· 1
Z 425 hCoV-19/Wales/PHWC-4A04E4/2020|EPI_ISL_743455|2020 ···················· ···················· ···················· ···················· NNNNNNNNNNNNNNNNNNNN NNNNNNNNNNNNNNNNNNNN 1
Z 426 hCoV-19/Wales/PHWC-4A04F3/2020|EPI_ISL_743456|2020 ···················· ···················· ···················· ···················· NNNNNNNNNNNNNNNNNNNN NNNNNNNNNNNN········ 1
Z 427 hCoV-19/Wales/PHWC-4A050F/2020|EPI_ISL_743457|2020 ···················· ···················· ···················· ···················· NNNNNNNNNNNNNNNNNNNN NNNNNNNNNNNNNNN····· 1
Z 513 hCoV-19/USA/CA-CZB-14232/2020|EPI_ISL_738900|2020- ···················· ···················· ···················· ···················· NNNNNNNNNNNNNNNNNNNN NNNNNNNNNNNNNNNNNNNN 1
Z 517 hCoV-19/USA/CA-CZB-14331/2020|EPI_ISL_738904|2020- ···················· ·A·················· ···················· ···················· NNNNNNNNNNNNNNNNNNNN NNNNNNNNNNNNNNNNNNNN 1
Z 519 hCoV-19/USA/CA-CZB-15305/2020|EPI_ISL_738906|2020- ···················· ·A·················· ···················· ···················· NNNNNNNNNNNNNNNNNNNN ··········NNNNN····· 1
Z 529 hCoV-19/USA/CA-CZB-14161/2020|EPI_ISL_738916|2020- ···················· ···················· ···················· ···················· NNNNNNNNNNNNNNNNNNNN NNNNNNNNNNNNNNN····· 1
Z 539 hCoV-19/USA/CA-CZB-14556/2020|EPI_ISL_738926|2020- ···················· ···················· ···················· ···················· NNNNNNNNNNNNNNNNNNNN NNNNNNNNNNNNNNNNNNNN 1
Z 540 hCoV-19/USA/CA-CZB-15257/2020|EPI_ISL_738927|2020- ···················· ···················· ···················· ···················· NNNNNNNNNNNNNNNNNNNN ···················· 1
Z 546 hCoV-19/USA/CA-CZB-15292/2020|EPI_ISL_738933|2020- ···················· ·A·················· ···················· ···················· NNNNNNNNNNNNNNNNNNNN NNNNNNNNNNNNNNNNNNNN 1
Z 553 hCoV-19/USA/CA-CZB-13786/2020|EPI_ISL_738940|2020- ···················· ···················· ···················· ···················· NNNNNNNNNNNNNNNNNNNN NNNNNNNNNNNNNNNNNNNN 1
Z 570 hCoV-19/USA/CA-CZB-13996/2020|EPI_ISL_738957|2020- ···················· ···················· ···················· ···················· NNNNNNNNNNNNNNNNNNNN NNNNNNNNNNNNNNNNNNNN 1
Z 573 hCoV-19/USA/CA-CZB-14584/2020|EPI_ISL_738960|2020- ···················· ···················· ···················· ···················· NNNNNNNNNNNNNNNNNNNN ···················· 1
Z 576 hCoV-19/USA/CA-CZB-13789/2020|EPI_ISL_738963|2020- ···················· ·A·················· ···················· ···················· NNNNNNNNNNNNNNNNNNNN ···················· 1
Z 579 hCoV-19/USA/CA-CZB-14650/2020|EPI_ISL_738966|2020- ···················· ···················· ···················· ···················· NNNNNNNNNNNNNNNNNNNN ···················· 1
Z 587 hCoV-19/USA/CA-CZB-13897/2020|EPI_ISL_738974|2020- ···················· ·A·················· ···················· ···················· NNNNNNNNNNNNNNNNNN·· ···················· 1
Z 589 hCoV-19/USA/CA-CZB-13948/2020|EPI_ISL_738976|2020- ···················· ···················· ···················· ···················· NNNNNNNNNNNNNNNNNNNN NNNNNNNNNNNNNNNNNNNN 1
Z 595 hCoV-19/USA/CA-CZB-13858/2020|EPI_ISL_738982|2020- ···················· ·A·················· ···················· ···················· NNNN················ ···················· 1
Z 609 hCoV-19/USA/CA-CZB-15338/2020|EPI_ISL_738996|2020- ···················· ·A·················· ···················· ···················· NNNNNNNNNNNNNNNNNNNN NNNNNNNNNNNNNNNNNNNN 1
Z 641 hCoV-19/Scotland/EDB10890/2020|EPI_ISL_742177|2020 ···················· ···················· ···················· ···················· NNNNNNNNNNNNNNNNNNNN NNNNNNNNNNNNNNNNNNNN 1
Z 644 hCoV-19/Scotland/EDB10908/2020|EPI_ISL_742183|2020 ···················· ···················· ···················· ···················· NNNNNNNNNNNNNNNNNNNN NNNNNNNNNNNNNNNNNNNN 1
Z 659 hCoV-19/Wales/PHWC-4A052D/2020|EPI_ISL_743459|2020 ···················· ···················· ···················· ···················· NNNNNNNNNNNNNNNNNNNN ···················· 1
Z 660 hCoV-19/Wales/PHWC-4A053C/2020|EPI_ISL_743460|2020 ···················· ···················· ···················· ···················· ······NNNNNNNNNNNNNN NNNNNNNNNNNNNNNNNNNN 1
Z 662 hCoV-19/Wales/PHWC-4A055A/2020|EPI_ISL_743462|2020 ···················· ···················· ···················· ···················· NNNNNNNNNNNNNNNNNNNN NNNNNNNNNNNNNNNNNNNN 1
Z 663 hCoV-19/Wales/PHWC-4A0578/2020|EPI_ISL_743463|2020 ···················· ···················· ···················· ···················· NNNNNNNNNNNNNNNNNNNN ···················· 1
Z 666 hCoV-19/Wales/PHWC-4A05E1/2020|EPI_ISL_743468|2020 ···················· ···················· ···················· ···················· NNNNNNNNNNNNNNNNNNNN NNNNNNNNNNNNNNNNNNNN 1
Z 668 hCoV-19/Wales/PHWC-4A061B/2020|EPI_ISL_743470|2020 ···················· ···················· ···················· ···················· NNNNNNNNNNNNNNNNNNNN ···················· 1
Z 669 hCoV-19/Wales/PHWC-4A062A/2020|EPI_ISL_743471|2020 ···················· ···················· ···················· ···················· ···················· NNNNNNNNN··········· 1
Z 672 hCoV-19/Wales/PHWC-4A0666/2020|EPI_ISL_743475|2020 ···················· ···················· ···················· ···················· NNNNNNNNNNNNNNNNNNNN ···················· 1
Z 673 hCoV-19/Wales/PHWC-4A0693/2020|EPI_ISL_743478|2020 ···················· ···················· ···················· ···················· ···················· NNNNNNNNNNNNNNNNNNNN 1
Z 674 hCoV-19/Wales/PHWC-4A06C0/2020|EPI_ISL_743481|2020 ···················· ···················· ···················· ···················· NNNNNNNNNNNNNNNNNNNN NNNNNNNNNNNNNNNNNNNN 1
Z 677 hCoV-19/Wales/PHWC-4A0709/2020|EPI_ISL_743485|2020 ···················· ···················· ···················· ···················· NNNNNNNNNNNNNNNNNNNN NNNNNNNNNNNNNNNNNNNN 1
Z 678 hCoV-19/Wales/PHWC-4A0718/2020|EPI_ISL_743486|2020 ···················· ···················· ···················· ···················· ···················· NNNNNNNNNNNNNNNNN··· 1
Z 679 hCoV-19/Wales/PHWC-4A0736/2020|EPI_ISL_743487|2020 ···················· ···················· ···················· ···················· NNNNNNNNNNNNNNNNNNNN ···················· 1
Z 682 hCoV-19/Wales/PHWC-4A0790/2020|EPI_ISL_743491|2020 ···················· ···················· ···················· ···················· NNNNNNNNNNNNNNNNNNNN NNN················· 1
Z 683 hCoV-19/Wales/PHWC-4A07BE/2020|EPI_ISL_743492|2020 ···················· ···················· ···················· ···················· NNNNNNNNNNNNNNNNNNNN NNNNNNNNNNNNNNNNNNNN 1
Z 687 hCoV-19/Wales/PHWC-4A09E5/2020|EPI_ISL_743526|2020 ···················· ···················· ···················· ···················· NNNNNNNNNNNNNNNNNNNN NNNNNNNNNNNNNNNNNNNN 1
Z 787 hCoV-19/Wales/PHWC-49AE67/2020|EPI_ISL_742262|2020 ···················· ·····A·············· ···················· ···················· NNNNNNNNNNNNNNNNNNNN NNNNNNNNNNNNNNNNNNNN 1
Z 789 hCoV-19/Wales/PHWC-49AE85/2020|EPI_ISL_742264|2020 ···················· ···················· ···················· ···················· NNNNNNNNNNNNNNNNNNNN NNNNNNNNNNNNNNNNNNNN 1
Z 792 hCoV-19/Wales/PHWC-49AEB2/2020|EPI_ISL_742267|2020 ···················· ···················· ···················· ···················· NNNNNNNNNNNNNNNNNNNN NNNNNNNNNN·········· 1
Z 793 hCoV-19/Wales/PHWC-49AEC1/2020|EPI_ISL_742268|2020 ···················· ···················· ···················· ···················· NNNNNNNNNNNNNNNNNNNN NNNNNNNNNNNNNNNNNNNN 1
Z 796 hCoV-19/Wales/PHWC-49AEFE/2020|EPI_ISL_742271|2020 ···················· ···················· ···················· ···················· NNNNNNNNNNNNNNNNNNNN NNNNNNNNNNNNNNNNNNNN 1
Z 804 hCoV-19/Wales/PHWC-49B060/2020|EPI_ISL_742287|2020 ···················· ···················· ···················· ···················· NNNNNNNNNNNNNNNNNNNN NNNNNNNNNNNNNNNNNNNN 1
Z 807 hCoV-19/Wales/PHWC-49B0BB/2020|EPI_ISL_742292|2020 ···················· ···················· ···················· ···················· NNNNNNNNNNNNNNNNN··· NNNNNNNNNNNNNNNNNNNN 1
Z 809 hCoV-19/Wales/PHWC-49B0D9/2020|EPI_ISL_742294|2020 ···················· ···················· ···················· ···················· NNNNNNNNNNNNNNNNNNNN NNNNNNNNNNNNNNNNNNNN 1
Z 810 hCoV-19/Wales/PHWC-49B0E8/2020|EPI_ISL_742295|2020 ···················· ···················· ···················· ···················· NNNNNNNNNNNNNNNNNNNN ···A················ 1
Z 811 hCoV-19/Wales/PHWC-49B0F7/2020|EPI_ISL_742296|2020 ···················· ···················· ···················· ···················· ···················· NNNNNNNNNNNNNNNNNNNN 1
Z 812 hCoV-19/Wales/PHWC-49B103/2020|EPI_ISL_742297|2020 ···················· ···················· ···················· ···················· NNNNNNNNNNNNNNNNNNNN NNNNNNNNNNNNNNNNNNNN 1
Z 814 hCoV-19/Wales/PHWC-49B121/2020|EPI_ISL_742299|2020 ···················· ···················· ···················· ···················· NNNNNNNNNNNNNNNNNNNN ···················· 1
Z 828 hCoV-19/Wales/PHWC-4A0833/2020|EPI_ISL_743500|2020 ···················· ···················· ···················· ···················· NNNNNNNNNNNNNNNNNNNN ···················· 1
Z 832 hCoV-19/Wales/PHWC-4A08D9/2020|EPI_ISL_743510|2020 ···················· ···················· ···················· ···················· NNNNNNNNNNNNNNNNNNNN NNNNNNNNNNNNNNN····· 1
Z 835 hCoV-19/Wales/PHWC-4A0912/2020|EPI_ISL_743513|2020 ···················· ···················· ···················· ···················· NNNNNNNNNNNNNNNNNNNN NNNNNNNNNNNNNNNNNNNN 1
Z 836 hCoV-19/Wales/PHWC-4A0930/2020|EPI_ISL_743515|2020 ···················· ···················· ···················· ···················· NNNN················ ···················· 1
Z 837 hCoV-19/Wales/PHWC-4A094F/2020|EPI_ISL_743516|2020 ···················· ···················· ···················· ···················· NNNN················ ···················· 1
Z 842 hCoV-19/Wales/PHWC-4A09C7/2020|EPI_ISL_743524|2020 ···················· ···················· ···················· ···················· NNNNNNNNNNNNNNNNNNNN NNNNNNNNNNNNNNNNNNNN 1
Z 872 hCoV-19/Wales/PHWC-4A0D16/2020|EPI_ISL_743568|2020 ···················· ···················· ···················· ···················· NNNNNNNNNNNNNNNNNNNN NNNNNNNNNNNNNNNNNNNN 1
Z 873 hCoV-19/Wales/PHWC-4A0D34/2020|EPI_ISL_743569|2020 ···················· ···················· ···················· ···················· NNNNNNNNNNNNNNNNNNNN NNNNNNNNNNNNNNNNNNNN 1
Z 877 hCoV-19/Wales/PHWC-4A0D8F/2020|EPI_ISL_743574|2020 ···················· ···················· ···················· ···················· ···················· ············NNN·NNNN 1
Z 879 hCoV-19/Wales/PHWC-4A0DAD/2020|EPI_ISL_743576|2020 ···················· ···················· ···················· ···················· NNNNNNNNNNNNNNNNNNNN NNNNNNNNNNNNNNNNNNNN 1
Z 882 hCoV-19/Wales/PHWC-4A0DF8/2020|EPI_ISL_743581|2020 ···················· ···················· ···················· ···················· NNNNNNNNNNNNNNNNNNNN NNNNNNNNNNNNNNN····· 1
Z 885 hCoV-19/Wales/PHWC-4A0E9B/2020|EPI_ISL_743587|2020 ···················· ···················· ···················· ···················· NNNNNNNNNNNNNNNNNNNN NNNNNNNNNNNNNNNNNNNN 1
Z 898 hCoV-19/Wales/PHWC-4A1049/2020|EPI_ISL_743611|2020 ···················· ···················· ···················· ···················· NNNNNNNNNNNNNNNNNNNN NNNNNNNNNNNNNNNNNNNN 1
Z 900 hCoV-19/Wales/PHWC-4A1243/2020|EPI_ISL_743642|2020 ···················· ···················· ···················· ···················· NNNNNNNNNNNNN······· ···················· 1
Z 901 hCoV-19/Wales/PHWC-4A1252/2020|EPI_ISL_743643|2020 ···················· ···················· ···················· ···················· ··NNN····NNNNNNNNNNN ···················· 1
Z 902 hCoV-19/Wales/PHWC-4A128F/2020|EPI_ISL_743646|2020 ···················· ···················· ···················· ···················· NNNNNNNNNNNNNNNNNNNN NNNNN··············· 1
Z 903 hCoV-19/Wales/PHWC-4A129E/2020|EPI_ISL_743647|2020 ···················· ···················· ···················· ···················· NNNNNNNNN··········· ···················· 1
Z 908 hCoV-19/Wales/PHWC-4A12F8/2020|EPI_ISL_743653|2020 ···················· ···················· ···················· ···················· NNNNNNNNNNNNNNNNNNNN ···················· 1
Z 1056 hCoV-19/England/CAMB-A97DB2/2020|EPI_ISL_741011|20 ···················· NNNNNNNNNNNNNNNNNNNN ···················· ···················· ···················· ···················· 1
Z 1155 hCoV-19/Wales/PHWC-49B561/2020|EPI_ISL_742352|2020 ···················· ···················· ···················· ···················· NNNNNNNNNNNNNNNNNNNN NNNNNNNNNNNNNNNNNNNN 1
Z 1156 hCoV-19/Wales/PHWC-49B59E/2020|EPI_ISL_742354|2020 ···················· ···················· ···················· ···················· NNNNNNNNNNNNNNNNNNNN NNNNNNNNNNNNNNNNNNNN 1
Z 1157 hCoV-19/Wales/PHWC-49B5BC/2020|EPI_ISL_742356|2020 ···················· ···················· ···················· ···················· ···················· ···········NNNN····· 1
Z 1158 hCoV-19/Wales/PHWC-49B5CB/2020|EPI_ISL_742357|2020 ···················· ···················· ···················· ···················· NNNNNNNNNNNNNNNNNNNN NNNNNNNNNNNNNNNNNNNN 1
Z 1160 hCoV-19/Wales/PHWC-49B5E9/2020|EPI_ISL_742359|2020 ···················· ···················· ···················· ···················· NNNNNNNNNNNNNNNNNNNN NNNNNNNNNNNNNNNNNNNN 1
```

| | | | | | | | | | |
|---|---|---|---|---|---|---|---|---|---|
| Z | 1161 | hCoV-19/Wales/PHWC-49B5F8/2020\|EPI_ISL_742360\|2020 | ···················· | ···················· | ···················· | ···················· | NNNNNNNNNNNNNNNNNNNN | NNNNNNNNNNNNNNNNNNNN | 1 |
| Z | 1162 | hCoV-19/Wales/PHWC-49B622/2020\|EPI_ISL_742361\|2020 | ···················· | ···················· | ···················· | ···················· | NNNNNNNNNNNNNNNNNNNN | NNNNNNNNNNNNNNNNNNNN | 1 |
| Z | 1163 | hCoV-19/Wales/PHWC-49B66E/2020\|EPI_ISL_742364\|2020 | ···················· | ···················· | ···················· | ···················· | NNNNNNNNNNNNNNNNNNNN | NNNNNNNNNNNNNNNNNNNN | 1 |
| Z | 1164 | hCoV-19/Wales/PHWC-49B67D/2020\|EPI_ISL_742365\|2020 | ···················· | ···················· | ···················· | ···················· | NNNNNNNNNNNNNNNNNNNN | ···················· | 1 |
| Z | 1166 | hCoV-19/Wales/PHWC-49B6AA/2020\|EPI_ISL_742368\|2020 | ···················· | ···················· | ···················· | ···················· | NNNNNNNNNNNNNNNNNNNN | NNNNNNNNNNNNNNNNNNNN | 1 |
| Z | 1168 | hCoV-19/Wales/PHWC-49B6C8/2020\|EPI_ISL_742370\|2020 | ···················· | ···················· | ···················· | ···················· | ···················· | NNNNNNNNNNNNNNN····· | 1 |
| Z | 1169 | hCoV-19/Wales/PHWC-49B6D7/2020\|EPI_ISL_742371\|2020 | ···················· | ···················· | ···················· | ···················· | NNNNNNNNNNNNNNNNNNNN | NNNNNNNNNNNNNNNNNNNN | 1 |
| Z | 1172 | hCoV-19/Wales/PHWC-49B701/2020\|EPI_ISL_742374\|2020 | ···················· | ···················· | ···················· | ···················· | NNNNNNNNNNNNNNNNNNNN | NNNNNNNNNNNNNNNNNNNN | 1 |
| Z | 1174 | hCoV-19/Wales/PHWC-49B74D/2020\|EPI_ISL_742378\|2020 | ···················· | ···················· | ···················· | ···················· | NNNNNNNNNNNNNNNNNNNN | NNNNNNNNNNNNNNNNNNNN | 1 |
| Z | 1176 | hCoV-19/Wales/PHWC-49B77A/2020\|EPI_ISL_742381\|2020 | ···················· | ···················· | ···················· | ···················· | ···················· | NNNNNNNNNNNNNNN····· | 1 |
| Z | 1177 | hCoV-19/Wales/PHWC-49B789/2020\|EPI_ISL_742382\|2020 | ···················· | ···················· | ···················· | ···················· | NNNNNNNNNNNNNNNNNNNN | NNNNNNNNNNNNNNNNNNNN | 1 |
| Z | 1178 | hCoV-19/Wales/PHWC-49B798/2020\|EPI_ISL_742383\|2020 | ···················· | ···················· | ···················· | ···················· | NNNNNNNNNNNNNNNNNNNN | NNNNNNNNNNNNNNNNNNNN | 1 |
| Z | 1179 | hCoV-19/Wales/PHWC-49B7D4/2020\|EPI_ISL_742386\|2020 | ···················· | ···················· | ···················· | ···················· | NNNNNNNNNNNNNNNNNNNN | NNNNNNNNNNNNNNNNNNNN | 1 |
| Z | 1180 | hCoV-19/Wales/PHWC-49B7E3/2020\|EPI_ISL_742387\|2020 | ···················· | ···················· | ···················· | ···················· | NNNNNNNNNNNNNNNNNNNN | NNNNNNNNNNNNNNNNNNNN | 1 |
| Z | 1181 | hCoV-19/Wales/PHWC-49B80E/2020\|EPI_ISL_742389\|2020 | ···················· | ···················· | ···················· | ···················· | NNNNNNNNNNNNNNNNNNNN | NNNNNNNNNNNNNNNNNNNN | 1 |
| Z | 1183 | hCoV-19/Wales/PHWC-49B859/2020\|EPI_ISL_742393\|2020 | ···················· | ···················· | ···················· | ···················· | NNNNNNNNNNNNNNNNNNNN | NNNNNNNNNNNNNNN····· | 1 |
| Z | 1184 | hCoV-19/Wales/PHWC-49B868/2020\|EPI_ISL_742394\|2020 | ···················· | ···················· | ···················· | ···················· | NNNNNNNNNNNN········ | ···················· | 1 |
| Z | 1185 | hCoV-19/Wales/PHWC-49B877/2020\|EPI_ISL_742395\|2020 | ···················· | ···················· | ···················· | ···················· | NNNNNNNNNNNNNNNNNNNN | ····NNNNNNN········· | 1 |
| Z | 1186 | hCoV-19/Wales/PHWC-49B886/2020\|EPI_ISL_742396\|2020 | ···················· | ···················· | ···················· | ···················· | NNNNNNNNNNNNNNNNN··· | ···················· | 1 |
| Z | 1187 | hCoV-19/Wales/PHWC-49B895/2020\|EPI_ISL_742397\|2020 | ···················· | ···················· | ···················· | ···················· | ···················· | NNNN·NNNNNNNNNN····· | 1 |
| Z | 1188 | hCoV-19/Wales/PHWC-49B8A4/2020\|EPI_ISL_742398\|2020 | ···················· | ···················· | ···················· | ···················· | NNNNNNNNNNNNNNNNNNNN | NNNNNNNNNNNNNNN····· | 1 |
| Z | 1189 | hCoV-19/Wales/PHWC-49B8B3/2020\|EPI_ISL_742399\|2020 | ···················· | ···················· | ···················· | ···················· | NNNNNNNNNNNNNNNNNNNN | NNNNNNNNNNNNNNNNNNNN | 1 |
| Z | 1190 | hCoV-19/Wales/PHWC-4A1067/2020\|EPI_ISL_743613\|2020 | ···················· | ···················· | ···················· | ···················· | NNNNNNNNNNNNNNNNN··· | ···················· | 1 |
| Z | 1191 | hCoV-19/Wales/PHWC-4A1155/2020\|EPI_ISL_743628\|2020 | ···················· | ···················· | ···················· | ···················· | NNNNNNNNNNNNNNNNNNNN | NNNNNNNNNNNNNNNNNNNN | 1 |
| Z | 1192 | hCoV-19/Wales/PHWC-4A1261/2020\|EPI_ISL_743644\|2020 | ···················· | ···················· | ···················· | ···················· | NNNNNNNNNNNNNNNNNNNN | NNNNN······NNNN····· | 1 |
| Z | 1193 | hCoV-19/Wales/PHWC-4A1270/2020\|EPI_ISL_743645\|2020 | ···················· | ···················· | ···················· | ···················· | NNNNNNNNNNNNNNNNNNNN | NNNNNNNNNNNNNNNNNNNN | 1 |
| Z | 1196 | hCoV-19/Wales/PHWC-4A135F/2020\|EPI_ISL_743659\|2020 | ···················· | ···················· | ···················· | ···················· | ···················· | NNNNNNNNN··········· | 1 |
| Z | 1199 | hCoV-19/Wales/PHWC-4A138C/2020\|EPI_ISL_743662\|2020 | ···················· | ···················· | ···················· | ···················· | NNNNNNNNNNNNNNNNNNNN | NNNNNNNNNNNNNNNNNNNN | 1 |
| Z | 1207 | hCoV-19/Wales/PHWC-4A145C/2020\|EPI_ISL_743674\|2020 | ···················· | ···················· | ···················· | ···················· | NNNNNNNNNNNNNNNNNNNN | ···············NNNNN | 1 |
| Z | 1211 | hCoV-19/Wales/PHWC-4A14A7/2020\|EPI_ISL_743679\|2020 | ···················· | ···················· | ···················· | ···················· | NNNNNNNNNNNNNNNNNNNN | ···················· | 1 |
| Z | 1216 | hCoV-19/Wales/PHWC-4A150E/2020\|EPI_ISL_743684\|2020 | ···················· | ···················· | ···················· | ···················· | NNNNNNNNNNNNNNNNNNNN | NNNNNNNNNNNNNNNNNNNN | 1 |
| Z | 1218 | hCoV-19/Wales/PHWC-4A152C/2020\|EPI_ISL_743686\|2020 | ···················· | ···················· | ···················· | ···················· | NNNNNNNNNNNN······NN | ···················· | 1 |
| Z | 1221 | hCoV-19/Wales/PHWC-4A1559/2020\|EPI_ISL_743689\|2020 | ···················· | ···················· | ···················· | ···················· | NNNN················ | ···················· | 1 |
| Z | 1225 | hCoV-19/Wales/PHWC-4A1595/2020\|EPI_ISL_743693\|2020 | ···················· | ······A············· | ···················· | ···················· | N······NNNNNNNNNNNNN | ···················· | 1 |
| Z | 1229 | hCoV-19/Wales/PHWC-4A15D1/2020\|EPI_ISL_743697\|2020 | ···················· | ···················· | ···················· | ···················· | NNNNNNNNNNNNNNNNNNNN | NNNNNNNNNNNNNNNNNNNN | 1 |
| Z | 1271 | hCoV-19/Italy/LAZ-INMI-119/2020\|EPI_ISL_746826\|202 | ···················· | ···················· | ···················· | ···················· | NNNNNNNNNNNNNNN····· | ···················· | 1 |
| Z | 1274 | hCoV-19/Italy/LAZ-INMI-122/2020\|EPI_ISL_746829\|202 | ···················· | ···················· | ···················· | ···················· | NNNNNNNNNNNNNN······ | ···················· | 1 |
| Z | 1325 | hCoV-19/Luxembourg/LNS0258260/2020\|EPI_ISL_739745\| | ···················· | ···A················ | ···················· | ···················· | NNNNNNNNNNNNNNNNNNNN | NNNNNNNNNNNNNNNNNNNN | 1 |
| Z | 1593 | hCoV-19/Wales/PHWC-49E5E6/2020\|EPI_ISL_743021\|2020 | ···················· | ···················· | ···················· | ···················· | NNNNNNNNNNNNNNNNNNNN | ···················· | 1 |
| Z | 1599 | hCoV-19/Wales/PHWC-49E65C/2020\|EPI_ISL_743028\|2020 | ···················· | ···················· | ···················· | ···················· | NNNNNNNNNNNNNNNNNNNN | ·················NNN | 1 |
| Z | 1604 | hCoV-19/Wales/PHWC-49E6A7/2020\|EPI_ISL_743033\|2020 | ···················· | ···················· | ···················· | ···················· | ··········NNNNNNNNNN | ···················· | 1 |
| Z | 2090 | hCoV-19/Wales/PHWC-49EC96/2020\|EPI_ISL_743108\|2020 | ···················· | ···················· | ···················· | ···················· | NNNNNNNNNNNNNNNNNNNN | NNNNNNNNNNNNNNNNNNNN | 1 |
| Z | 2093 | hCoV-19/Wales/PHWC-49ECF0/2020\|EPI_ISL_743111\|2020 | ···················· | ···················· | ···················· | ···················· | NNNNNNNNNNNNNNNNNNNN | NNNNNNNNNNNNNNNNNNNN | 1 |
| Z | 2098 | hCoV-19/Wales/PHWC-49ED57/2020\|EPI_ISL_743116\|2020 | ···················· | ···················· | ···················· | ···················· | NNNNNNNNNNNNNNNNNNNN | ···················· | 1 |
| Z | 2099 | hCoV-19/Wales/PHWC-49ED66/2020\|EPI_ISL_743117\|2020 | ···················· | ···················· | ···················· | ···················· | NNNNNNNNNNNNNNNNNNNN | NNNNNNNNNNNNNNNNNNNN | 1 |
| Z | 2101 | hCoV-19/Wales/PHWC-49ED84/2020\|EPI_ISL_743119\|2020 | ···················· | ···················· | ···················· | ···················· | NNNNNNNNNNNNNNNNNNNN | NNNNNNNNNNNNNNNNNNNN | 1 |
| Z | 2104 | hCoV-19/Wales/PHWC-49EDC0/2020\|EPI_ISL_743123\|2020 | ···················· | ···················· | ···················· | ···················· | NNNNNNNNNNNNNNNNNNNN | NNNNNNNNNNNNNNNNNNNN | 1 |
| Z | 2105 | hCoV-19/Wales/PHWC-49EDDF/2020\|EPI_ISL_743124\|2020 | ···················· | ···················· | ···················· | ···················· | NNNNNNNNNNNNNNNNNNNN | NNNNNNNNNNNNNNNNNNNN | 1 |
| Z | 2107 | hCoV-19/Wales/PHWC-49EDFD/2020\|EPI_ISL_743126\|2020 | ···················· | ···················· | ···················· | ···················· | NNNNNNNNNNNNNNNNNNNN | NNNNNNNNNNNNNNNNNNNN | 1 |
| Z | 2109 | hCoV-19/Wales/PHWC-49EE27/2020\|EPI_ISL_743129\|2020 | ···················· | ···················· | ···················· | ···················· | ···················· | NNNNN··············· | 1 |
| Z | 2111 | hCoV-19/Wales/PHWC-49EE63/2020\|EPI_ISL_743133\|2020 | ···················· | ···················· | ···················· | ···················· | NNNNNNNNNNNNNNNNNNNN | ···················· | 1 |
| Z | 2112 | hCoV-19/Wales/PHWC-49EE72/2020\|EPI_ISL_743134\|2020 | ···················· | ···················· | ···················· | ···················· | NNNNNNNNNNNNNNNNNNNN | NN·················· | 1 |
| Z | 2114 | hCoV-19/Wales/PHWC-49EE90/2020\|EPI_ISL_743136\|2020 | ···················· | ···················· | ···················· | ···················· | NNNNNNNNNNNNNNNNNNNN | ···················· | 1 |
| Z | 2123 | hCoV-19/Wales/PHWC-49EF51/2020\|EPI_ISL_743147\|2020 | ···················· | ···················· | ···················· | ···················· | NNNNNNNNNNNNNNNNNNNN | NNNNNNNNNNNNNNNNNNNN | 1 |
| Z | 2127 | hCoV-19/Wales/PHWC-49EFCA/2020\|EPI_ISL_743152\|2020 | ···················· | ···················· | ···················· | ···················· | NNNNNNNNNNNN········ | NNNNNNNNNNNNNNNNNNNN | 1 |
| Z | 2128 | hCoV-19/Wales/PHWC-49EFD9/2020\|EPI_ISL_743153\|2020 | ···················· | ···················· | ···················· | ···················· | NNNNNNNNNNNNNNNNNNNN | ···················· | 1 |
| Z | 2129 | hCoV-19/Wales/PHWC-49EFF7/2020\|EPI_ISL_743155\|2020 | ···················· | ···················· | ···················· | ···················· | NNNNNNNNNNNNNNNNNNNN | NNNNNNNNNNNNNNNNNNNN | 1 |
| Z | 2130 | hCoV-19/Wales/PHWC-49F011/2020\|EPI_ISL_743157\|2020 | ···················· | ···················· | ···················· | ···················· | NNNN················ | ···················· | 1 |
| Z | 2137 | hCoV-19/Wales/PHWC-49F0D5/2020\|EPI_ISL_743166\|2020 | ···················· | ···················· | ···················· | ···················· | NNNNNNNNNNNNNNNNNNNN | NNNNNNNNNNNNNNNNNNNN | 1 |
| Z | 2138 | hCoV-19/Wales/PHWC-49F10F/2020\|EPI_ISL_743167\|2020 | ···················· | ···················· | ···················· | ···················· | NNNNNNNNNNNNNNNNNNNN | ···················· | 1 |
| Z | 2139 | hCoV-19/Wales/PHWC-49F11E/2020\|EPI_ISL_743168\|2020 | ···················· | ···················· | ···················· | ···················· | NNNNNNNNNNNNNNNNNNNN | NNNNNNNNNNNNNNNNNNNN | 1 |
| Z | 2141 | hCoV-19/Wales/PHWC-49F14B/2020\|EPI_ISL_743171\|2020 | ···················· | ···················· | ···················· | ···················· | NNNNNNNNNNNNNNNNNNNN | NNNNNNNNNNNNNNNNNNNN | 1 |
| Z | 2143 | hCoV-19/Wales/PHWC-49F169/2020\|EPI_ISL_743173\|2020 | ···················· | ···················· | ···················· | ···················· | NNNNNNNNNNNNNNNNNNNN | ···················· | 1 |
| Z | 2148 | hCoV-19/Wales/PHWC-49F1D2/2020\|EPI_ISL_743178\|2020 | ···················· | ···················· | ···················· | ···················· | NNNN·········NNNNNNN | ···················· | 1 |
| Z | 2305 | hCoV-19/USA/CA-CZB-14008/2020\|EPI_ISL_738678\|2020- | ···················· | ···················· | ···················· | ···················· | ·NNNNNNNNNNNNNNNNNNN | ···················· | 1 |
| Z | 2424 | hCoV-19/England/CAMB-A97B12/2020\|EPI_ISL_740999\|20 | ···················· | NNNNNNNNNNNNNNNNNNNN | ···················· | ···················· | ···················· | ···················· | 1 |
| Z | 2425 | hCoV-19/Wales/PHWC-49F1F0/2020\|EPI_ISL_743180\|2020 | ···················· | ···················· | ···················· | ···················· | NNNNNNNNNNNNNNNNNNNN | ···················· | 1 |
| Z | 2426 | hCoV-19/Wales/PHWC-49F22A/2020\|EPI_ISL_743182\|2020 | ···················· | ···················· | ···················· | ···················· | ···················· | NNNNNNN············· | 1 |
| Z | 2427 | hCoV-19/Wales/PHWC-49F248/2020\|EPI_ISL_743184\|2020 | ···················· | ···················· | ···················· | ···················· | NNNNNNNNNNNNNNNNNNNN | NNNNNNNNNNNNNNNNNNNN | 1 |
| Z | 2429 | hCoV-19/Wales/PHWC-49F284/2020\|EPI_ISL_743188\|2020 | ···················· | ···················· | ···················· | ···················· | NNNNNNNNNNNNNNNNNNNN | ···NNNNNNNNNNNN····· | 1 |
| Z | 2434 | hCoV-19/Wales/PHWC-49F2FD/2020\|EPI_ISL_743195\|2020 | ···················· | ···················· | ···················· | ···················· | NNNNNNNNNNNNNNNNNNNN | NNNNNNNNNNNNNNNNNNNN | 1 |
| Z | 2435 | hCoV-19/Wales/PHWC-49F309/2020\|EPI_ISL_743196\|2020 | ···················· | ···················· | ···················· | ···················· | NNNNNNNNNNNNNNNNNNNN | ···················· | 1 |
| Z | 2437 | hCoV-19/Wales/PHWC-49F336/2020\|EPI_ISL_743199\|2020 | ···················· | ···················· | ···················· | ···················· | NNNNNNNNNNNNNNNNNNNN | NNNNN··············· | 1 |
| Z | 2439 | hCoV-19/Wales/PHWC-49F372/2020\|EPI_ISL_743203\|2020 | ···················· | ···················· | ···················· | ···················· | NNNNNNNNNNNNNNNNNNNN | NNNNNNNNNNNNNNNNNNNN | 1 |
| Z | 2444 | hCoV-19/Wales/PHWC-49F3CD/2020\|EPI_ISL_743208\|2020 | ···················· | ···················· | ···················· | ···················· | ···················· | NNNNNNNNNNNNNNNNNNNN | 1 |
| Z | 2574 | hCoV-19/USA/CA-CZB-14540/2020\|EPI_ISL_738726\|2020- | ···················· | ·A·················· | ···················· | ···················· | NNNNNNNNNNNNNNNNNNNN | NNNNNNNNNNNNNNNNNNNN | 1 |
| Z | 2580 | hCoV-19/USA/CA-CZB-13828/2020\|EPI_ISL_738743\|2020- | ···················· | ···················· | ···················· | ···················· | NNNNNNNNNNNNNNNNNNNN | NNNNNNNNNNNNNNNNNNNN | 1 |
| Z | 2582 | hCoV-19/USA/CA-CZB-13738/2020\|EPI_ISL_738745\|2020- | ···················· | ···················· | ···················· | ···················· | NNNNNNNNNNN········· | ···················· | 1 |
| Z | 2585 | hCoV-19/USA/CA-CZB-13919/2020\|EPI_ISL_738748\|2020- | ···················· | ·A·················· | ···················· | ···················· | NNNNNNNNNNNNNNNNNNNN | NNNNNNNNNNNNNNN····· | 1 |
| Z | 2586 | hCoV-19/USA/CA-CZB-13986/2020\|EPI_ISL_738749\|2020- | ···················· | ···················· | ···················· | ···················· | ···················· | NNNNNNNNNNNNNNNNNNNN | 1 |

| | | | | | | | | | |
|---|---|---|---|---|---|---|---|---|---|
| Z | 2599 | hCoV-19/USA/CA-CZB-14221/2020\|EPI_ISL_738762\|2020- | ···················· | ·A·················· | ···················· | ···················· | NNNNNNNNNNNNNNNNNNNN | ···················· | 1 |
| Z | 2602 | hCoV-19/USA/CA-CZB-14533/2020\|EPI_ISL_738765\|2020- | ···················· | ·A·················· | ···················· | ···················· | NNNNNNNNNNNNNNNNNNNN | NNNNNNNNNNNNNNNNNNNN | 1 |
| Z | 2603 | hCoV-19/USA/CA-CZB-13909/2020\|EPI_ISL_738766\|2020- | ···················· | ···················· | ···················· | ···················· | NNNN················ | ···················· | 1 |
| Z | 2604 | hCoV-19/USA/CA-CZB-13777/2020\|EPI_ISL_738767\|2020- | ···················· | ·A·················· | ···················· | ···················· | NNNNNNNNNNNNNNNNNNNN | NNNNNNNNNNNNNNNNNNNN | 1 |
| Z | 2605 | hCoV-19/USA/CA-CZB-14109/2020\|EPI_ISL_738768\|2020- | ···················· | ···················· | ···················· | ···················· | NNNNNNNNNNNNNNNNNNNN | NNNNNNNNNNNNNNNNNNNN | 1 |
| Z | 2611 | hCoV-19/USA/CA-CZB-14261/2020\|EPI_ISL_738774\|2020- | ···················· | ···················· | ···················· | ···················· | NNNNNNNNNNNNNNNNNNNN | NNNNNNNNNNNNNNNNNNNN | 1 |
| Z | 2612 | hCoV-19/USA/CA-CZB-13949/2020\|EPI_ISL_738775\|2020- | ···················· | ···················· | ···················· | ···················· | NNNNNNNNNNNNNNNNNNNN | NNNNNNNNNNNNNNNNNNNN | 1 |
| Z | 2618 | hCoV-19/USA/CA-CZB-14373/2020\|EPI_ISL_738781\|2020- | ···················· | ···················· | ···················· | ···················· | NNNNNNNNNNNNNNNNNNNN | NNNNNNNNNNNNNNNNNNNN | 1 |
| Z | 2629 | hCoV-19/USA/CA-CZB-13774/2020\|EPI_ISL_738792\|2020- | ···················· | ·A·················· | ···················· | ···················· | NNNNNNNNNNNNNNNNNNNN | NNNNNNNNNNNNNNNNNNNN | 1 |
| Z | 2630 | hCoV-19/USA/CA-CZB-13819/2020\|EPI_ISL_738793\|2020- | ···················· | ·A·················· | ···················· | ···················· | NNNNNNNNNNNNNNNNNNNN | ·······NNNNNNNNNNNNN | 1 |
| Z | 2631 | hCoV-19/USA/CA-CZB-15293/2020\|EPI_ISL_738794\|2020- | ···················· | ·A·················· | ···················· | ···················· | NNNNNNNNNNNNNNNNNNNN | NNNNNNNNNNNNNNNNNNNN | 1 |
| Z | 2633 | hCoV-19/USA/CA-CZB-13787/2020\|EPI_ISL_738796\|2020- | ···················· | ···················· | ···················· | ···················· | NNNNNNNNNNNNNNNNNNNN | NNNNNNNNNNNNNNNNNNNN | 1 |
| Z | 2635 | hCoV-19/USA/CA-CZB-14570/2020\|EPI_ISL_738798\|2020- | ···················· | ···················· | ···················· | ···················· | NNNNNNNNNNNNNNNNNNNN | NNNNNNNNNNNNNNN····M | 1 |
| Z | 2637 | hCoV-19/USA/CA-CZB-15170/2020\|EPI_ISL_739234\|2020- | ···················· | ···················· | ···················· | ···················· | NNNNNNNNNNNNNNNNNNNN | NNNNNNNNNNNNNNNNNNNN | 1 |
| Z | 2639 | hCoV-19/USA/CA-CZB-14387/2020\|EPI_ISL_739236\|2020- | ···················· | ···················· | ···················· | ···················· | NNNN················ | NNNNNNNNNNNNNNNNNNNN | 1 |
| Z | 2662 | hCoV-19/Wales/PHWC-49DB12/2020\|EPI_ISL_742876\|2020 | ···················· | ···················· | ···················· | ···················· | NNNNNNNNNNNNNNNNNNNN | NNNNNNNNNNNNNNNNNNNN | 1 |
| Z | 2668 | hCoV-19/Wales/PHWC-49DBF4/2020\|EPI_ISL_742884\|2020 | ···················· | ···················· | ···················· | ···················· | ···················· | ··········NNNN······ | 1 |
| Z | 2669 | hCoV-19/Wales/PHWC-49DC00/2020\|EPI_ISL_742885\|2020 | ···················· | ···················· | ···················· | ···················· | NNNNNNNNNNNNNNNNNNNN | NNNNNNNNNNNNNNNNNNNN | 1 |
| Z | 2677 | hCoV-19/Wales/PHWC-49F4BB/2020\|EPI_ISL_743223\|2020 | ···················· | ···················· | ···················· | ···················· | NNNNNNNNNNNNNNNNNNNN | NNNNNNNNNNNNNNN····· | 1 |
| Z | 2686 | hCoV-19/Wales/PHWC-49F56D/2020\|EPI_ISL_743233\|2020 | ···················· | ···················· | ···················· | ···················· | NNNNNNNNNNNNNNNNNNNN | ···················· | 1 |
| Z | 2688 | hCoV-19/Wales/PHWC-49F59A/2020\|EPI_ISL_743235\|2020 | ···················· | ···················· | ···················· | ···················· | NNNNNNNNNNNNNNNNNNNN | ···················· | 1 |
| Z | 2692 | hCoV-19/Wales/PHWC-49F5E5/2020\|EPI_ISL_743240\|2020 | ···················· | ···················· | ···················· | ···················· | NNNNNNNNNNNNNNNNNN·· | ···················· | 1 |
| Z | 2696 | hCoV-19/Wales/PHWC-49F66A/2020\|EPI_ISL_743247\|2020 | ···················· | ···················· | ···················· | ···················· | NNNNNNNNNNNNNNNNNNNN | ···················· | 1 |
| Z | 2817 | hCoV-19/USA/CA-CZB-14356/2020\|EPI_ISL_739200\|2020- | ···················· | ·············A······ | ···················· | ···················· | NNNNNNNNNNNNNNNNNNNN | NNNNNNNNNNNNNNNNNNNN | 1 |
| Z | 2824 | hCoV-19/USA/CA-CZB-14117/2020\|EPI_ISL_739207\|2020- | ···················· | ···················· | ···················· | ···················· | NNNNNNNNNNNNNNNNNNNN | NNNNNNNNNNNNNNNNNNNN | 1 |
| Z | 2829 | hCoV-19/USA/CA-CZB-14380/2020\|EPI_ISL_739212\|2020- | ···················· | ·A·················· | ···················· | ···················· | NNNNNNNNNNNNNNNNNNNN | NNNNNNNNNNNNNNNNNNNN | 1 |
| Z | 2832 | hCoV-19/USA/CA-CZB-14426/2020\|EPI_ISL_739215\|2020- | ···················· | ···················· | ···················· | ···················· | NNNNNNNNNNNNNNNNNNNN | NNNNNNNNNNNNNNNNNNNN | 1 |
| Z | 2836 | hCoV-19/USA/CA-CZB-15178/2020\|EPI_ISL_739219\|2020- | ···················· | ·A·················· | ···················· | ···················· | NNNNNNNNNNNNNNNNNNNN | ···················· | 1 |
| Z | 2844 | hCoV-19/USA/CA-CZB-14657/2020\|EPI_ISL_739227\|2020- | ···················· | ·A·················· | ···················· | ···················· | NNNNNNNNNNNNNNNNNNNN | ···················· | 1 |
| Z | 2849 | hCoV-19/USA/CA-CZB-14986/2020\|EPI_ISL_739232\|2020- | ···················· | ·A·················· | ···················· | ···················· | NNNNNNNNNNNNNNNNNNNN | NNNNNNNNNNNNNNNNNNNN | 1 |
| Z | 2857 | hCoV-19/USA/CA-CZB-15009/2020\|EPI_ISL_739246\|2020- | ···················· | ·A·················· | ···················· | ···················· | NNNNNNNNNNNNNNNNNNNN | ···················· | 1 |
| Z | 2858 | hCoV-19/USA/CA-CZB-15258/2020\|EPI_ISL_739247\|2020- | ···················· | ···················· | ···················· | ···················· | NNNNNNNNNNNNNNNNNNNN | ·············M······ | 1 |
| Z | 2859 | hCoV-19/USA/CA-CZB-15321/2020\|EPI_ISL_739248\|2020- | ···················· | ·A·················· | ···················· | ···················· | NNNNNNNNNNNNNNNNNNNN | NNNNNNNNNNNNNNNNNNNN | 1 |
| Z | 2866 | hCoV-19/USA/CA-CZB-14651/2020\|EPI_ISL_739255\|2020- | ···················· | ···················· | ···················· | ···················· | NNNNNNNNNNNNNNNNNNNN | NNNNNNNNNNNNNNNNNNNN | 1 |
| Z | 2867 | hCoV-19/USA/CA-CZB-13697/2020\|EPI_ISL_739256\|2020- | ···················· | ·A·················· | ···················· | ···················· | NNNNNNNNNNNNNNNNNNNN | NNNNNNNNNNNNNNNNNNNN | 1 |
| Z | 2871 | hCoV-19/USA/CA-CZB-13700/2020\|EPI_ISL_739260\|2020- | ···················· | ···················· | ···················· | ···················· | NNNNNNNNNNNNNNNN···· | ···················· | 1 |
| Z | 2878 | hCoV-19/USA/CA-CZB-14908/2020\|EPI_ISL_739267\|2020- | ···················· | ···················· | ···················· | ···················· | NNNNNNNNNNNNNNNNNNNN | NNNNNNNNNNNNNNN····· | 1 |
| Z | 2889 | hCoV-19/USA/CA-CZB-15297/2020\|EPI_ISL_739278\|2020- | ···················· | ·A·················· | ···················· | ···················· | NNNNNNNNNNNNNNNNNNNN | NNNNNNNNNNNNNNNNNNNN | 1 |
| Z | 2894 | hCoV-19/USA/CA-CZB-13772/2020\|EPI_ISL_739283\|2020- | ···················· | ·A·················· | ···················· | ···················· | NNNNNNNNNNNNNNNNNNNN | ·········NNNNNNNNNNN | 1 |
| Z | 2897 | hCoV-19/USA/CA-CZB-14135/2020\|EPI_ISL_739286\|2020- | ···················· | ···················· | ···················· | ···················· | NNNNNNNNNNNNNNNNNNNN | ···················· | 1 |
| Z | 2901 | hCoV-19/USA/CA-CZB-14085/2020\|EPI_ISL_739290\|2020- | ···················· | ···················· | ···················· | ···················· | NNNNNNNNNNNNNNNNNNNN | ···················· | 1 |
| Z | 2905 | hCoV-19/USA/CA-CZB-14000/2020\|EPI_ISL_739294\|2020- | ···················· | ···················· | ···················· | ···················· | ···················· | NNNNNNNNNNNNNNNNN··· | 1 |
| Z | 3039 | hCoV-19/Wales/PHWC-49D972/2020\|EPI_ISL_742854\|2020 | ···················· | ···················· | ···················· | ···················· | NNNNNNNNNNNNNNNNNNNN | NNNNNNNNNNNNNNNNNNNN | 1 |
| Z | 3047 | hCoV-19/Wales/PHWC-49DA15/2020\|EPI_ISL_742863\|2020 | ···················· | ···················· | ···················· | ···················· | NNNNNNNNNNNNNNNNNNNN | ···················· | 1 |
| Z | 3059 | hCoV-19/Wales/PHWC-49DC97/2020\|EPI_ISL_742893\|2020 | ···················· | ···················· | ···················· | ···················· | NNNNNNNNNNNNNNNNNNNN | NNNNNNNNNNNNNNNNNNNN | 1 |
| Z | 3062 | hCoV-19/Wales/PHWC-49DD0D/2020\|EPI_ISL_742899\|2020 | ···················· | ···················· | ···················· | ···················· | NNNNNNNNNNNN········ | NNNNNNNNNNNNNNNNNNNN | 1 |
| Z | 3075 | hCoV-19/USA/CA-CZB-13891/2020\|EPI_ISL_739302\|2020- | ···················· | ·A·················· | ···················· | ···················· | NNNNNNNNNNNNNNNNNNNN | NNNR················ | 1 |
| Z | 3076 | hCoV-19/USA/CA-CZB-13814/2020\|EPI_ISL_739303\|2020- | ···················· | ···················· | ···················· | ···················· | NNNNNNNNNNNNNNNNNNNN | NNNNNNNNNNNNNNNNNNNN | 1 |
| Z | 3090 | hCoV-19/USA/CA-CZB-15316/2020\|EPI_ISL_739317\|2020- | ···················· | ·A·················· | ···················· | ···················· | NNNNNNNNNNNNNNNNNNNN | NNNNNNNNNNNNNNNNNNNN | 1 |
| Z | 3094 | hCoV-19/USA/CA-CZB-14063/2020\|EPI_ISL_739337\|2020- | ···················· | ·R·················· | ···················· | ···················· | NNNNNNNNNNNNNNNNNNNN | NNNNNNNNNNNNNNNNNNNN | 1 |
| Z | 3106 | hCoV-19/USA/CA-CZB-14360/2020\|EPI_ISL_739349\|2020- | ···················· | ···A················ | ···················· | ···················· | NNNNNNNNNNNNNNNNNNNN | NNNNNNNNNNNNNNNNNNNN | 1 |
| Z | 3109 | hCoV-19/USA/CA-CZB-14996/2020\|EPI_ISL_739352\|2020- | ···················· | ···················· | ···················· | ···················· | NNNNNNNNNNNNNNNNNNNN | NNNNNNNNNNNNNNNNNNNN | 1 |
| Z | 3113 | hCoV-19/USA/CA-CZB-13868/2020\|EPI_ISL_739356\|2020- | ···················· | ·A·················· | ···················· | ···················· | NNNNNNNNNNNNNNNNNNNN | NNNNNNNNNNNNNNNNNNNN | 1 |
| Z | 3114 | hCoV-19/USA/CA-CZB-14566/2020\|EPI_ISL_739357\|2020- | ···················· | ···················· | ···················· | ···················· | NNNNNNNNNNNNNNNNNNNN | NNNNNNNNNNNNNNNNNNNN | 1 |
| Z | 3176 | hCoV-19/Wales/PHWC-49DD49/2020\|EPI_ISL_742903\|2020 | ···················· | ···················· | ···················· | ···················· | NNNNNNNNNNNNNNNNNNNN | NNNNNNNNNNNNNNNNNNNN | 1 |
| Z | 3187 | hCoV-19/Wales/PHWC-49DE0A/2020\|EPI_ISL_742915\|2020 | ···················· | ······A············· | ···················· | ···················· | NNNNNNNNNNN········· | ···················· | 1 |
| Z | 3230 | hCoV-19/Wales/PHWC-49E13D/2020\|EPI_ISL_742958\|2020 | ···················· | ···················· | ···················· | ···················· | NNNNNNNNNNNNNNNNNNNN | ···················· | 1 |
| Z | 3244 | hCoV-19/Wales/PHWC-49E337/2020\|EPI_ISL_742988\|2020 | ···················· | ···················· | ···················· | ···················· | NNNNN··············· | ···················· | 1 |
| Z | 3332 | hCoV-19/USA/CA-CZB-14420/2020\|EPI_ISL_739323\|2020- | ···················· | ·A·················· | ···················· | ···················· | NNNNNNNNNNNNNNNNNNNN | NNNNNNNNNNNNNNNNNNNN | 1 |
| Z | 3337 | hCoV-19/USA/CA-CZB-14245/2020\|EPI_ISL_739328\|2020- | ···················· | ···················· | ···················· | ···················· | NNNNNNNNNNNNNNNNNNNN | NNNNNNNNNNNNNNNNNNNN | 1 |
| Z | 3361 | hCoV-19/USA/CA-CZB-15023/2020\|EPI_ISL_739376\|2020- | ···················· | ·A·················· | ···················· | ···················· | NNNNNNNNNNNNNNNNNNNN | NNNNNNNNNNNNNNNNNNNN | 1 |
| Z | 3377 | hCoV-19/USA/CA-CZB-15328/2020\|EPI_ISL_739392\|2020- | ···················· | ·A·················· | ···················· | ···················· | NNNNNNNNNNNNNNNNNNNN | NNNNNNNNNNNNNNNNNNNN | 1 |
| Z | 3479 | hCoV-19/Belgium/UGent-316/2020\|EPI_ISL_745205\|2020 | ···················· | ···················· | ···················· | ···················· | NNNNNNNNNNNNNNNNNNNN | NNNNNNNNNNNNNNNNNNNN | 1 |
| Z | 3482 | hCoV-19/Belgium/UGent-322/2020\|EPI_ISL_745209\|2020 | ···················· | ···················· | ···················· | ···················· | NNNNNNNNNNNNNNNNNNNN | NNNNNNNNNNNNNNNNNNNN | 1 |
| Z | 3483 | hCoV-19/Belgium/UGent-324/2020\|EPI_ISL_745211\|2020 | ···················· | ···················· | ···················· | ···················· | NNNNNNNNNNNNNNNNNNNN | NNNNNNNNNNNNNNNNNNNN | 1 |
| Z | 3484 | hCoV-19/Belgium/UGent-325/2020\|EPI_ISL_745212\|2020 | ···················· | ···················· | ···················· | ···················· | NNNNNNNNNNNNNNNNNNNN | NNNNNNNNNNNNNNNNNNNN | 1 |
| Z | 3489 | hCoV-19/Belgium/UGent-333/2020\|EPI_ISL_745218\|2020 | ···················· | ···················· | ···················· | ···················· | NNNNNNNNNNNNNNNNNNNN | NNNNNNNNNNNNNNNNNNNN | 1 |
| Z | 3490 | hCoV-19/Belgium/UGent-334/2020\|EPI_ISL_745219\|2020 | ···················· | ···················· | ···················· | ···················· | NNNNNNNNNNNNNNNNNNNN | NNNNNNNNNNNNNNNNNNNN | 1 |
| Z | 3592 | hCoV-19/USA/CA-CZB-14334/2020\|EPI_ISL_739407\|2020- | ···················· | ·A·················· | ···················· | ···················· | NNNNNNNNNNNNNNNNNNNN | NNNNNNNNNNNNNNNNNNNN | 1 |
| Z | 3609 | hCoV-19/USA/CA-CZB-14112/2020\|EPI_ISL_739424\|2020- | ···················· | ···················· | ···················· | ···················· | NNNNNNNNNNNNNNNNNNNN | NNNNNNNNNNNNNNNNNNNN | 1 |
| Z | 3611 | hCoV-19/USA/CA-CZB-14279/2020\|EPI_ISL_739426\|2020- | ···················· | ·A·················· | ···················· | ···················· | NNNNNNNNNNNNNNNNNNNN | NNNNNNNNNNNNNNNNNNNN | 1 |
| Z | 3616 | hCoV-19/USA/CA-CZB-15299/2020\|EPI_ISL_739431\|2020- | ···················· | ·A·················· | ···················· | ···················· | NNNNNNNNNNNNNNNNNNNN | NNNNNNNNNNNNNNNNNNNN | 1 |
| Z | 3628 | hCoV-19/USA/CA-CZB-13806/2020\|EPI_ISL_739448\|2020- | ···················· | ···················· | ···················· | ···················· | NNNNNNNNNNNN········ | ···················· | 1 |
| Z | 3629 | hCoV-19/USA/CA-CZB-13902/2020\|EPI_ISL_739449\|2020- | ···················· | ·A·················· | ···················· | ···················· | NNNNNNNNNNNNNNNNNNNN | NNNNNNNNNNNNNNNNNNNN | 1 |
| Z | 3633 | hCoV-19/USA/CA-CZB-14578/2020\|EPI_ISL_739453\|2020- | ···················· | ···················· | ···················· | ···················· | NNNNNNNNNNNNNNNNNNNN | NNNNNNNNNNNNNNNNNNNN | 1 |
| Z | 3744 | hCoV-19/England/PORT-2F62AB/2020\|EPI_ISL_741756\|20 | ···················· | ···················· | ···················· | ···················· | NNNNNNNNNNNNNNNNNNNN | NNNNNNNNNNNNNNNNNNNN | 1 |
| Z | 3841 | hCoV-19/USA/CA-CZB-15272/2020\|EPI_ISL_739440\|2020- | ···················· | ···················· | ···················· | ···················· | NNNNNNNNNNNNNNNNNNNN | NNNNNNNNNNNNNNNNNNNN | 1 |
| Z | 3845 | hCoV-19/USA/CA-CZB-13741/2020\|EPI_ISL_739444\|2020- | ···················· | ·A·················· | ···················· | ···················· | ···················· | NNNNNNN············· | 1 |
| Z | 3847 | hCoV-19/USA/CA-CZB-15036/2020\|EPI_ISL_739481\|2020- | ···················· | ·A·················· | ···················· | ···················· | NNNNNNNNNNNNNNNNNNNN | ···················· | 1 |
| Z | 3849 | hCoV-19/USA/CA-CZB-14265/2020\|EPI_ISL_739483\|2020- | ···················· | ···················· | ···················· | ···················· | NNNNNNNNNNNNNN······ | ···················· | 1 |

<table>
<tr><td>Z</td><td>3861</td><td>hCoV-19/USA/CA-CZB-14084/2020|EPI_ISL_739495|2020-</td><td>····················</td><td>····················</td><td>····················</td><td>····················</td><td>NNNNNNNNNNNNNNNNNNNN</td><td>····················</td><td>1</td></tr>
<tr><td>Z</td><td>3903</td><td>hCoV-19/Wales/PHWC-4A2C80/2020|EPI_ISL_744000|2020</td><td>····················</td><td>····················</td><td>····················</td><td>····················</td><td>NNNNNNNNNNNNNNNNNNNN</td><td>····················</td><td>1</td></tr>
<tr><td>Z</td><td>3906</td><td>hCoV-19/Wales/PHWC-4A2CBD/2020|EPI_ISL_744003|2020</td><td>····················</td><td>····················</td><td>····················</td><td>····················</td><td>NNNNNNNNNNNNNNNNNNNN</td><td>NNNNNNNNNNNNNNNNNNNN</td><td>1</td></tr>
<tr><td>Z</td><td>3918</td><td>hCoV-19/Wales/PHWC-4A2DAB/2020|EPI_ISL_744016|2020</td><td>····················</td><td>····················</td><td>····················</td><td>····················</td><td>NNNNNNNNNNNNNNNNNNNN</td><td>NNNNNNNNNNNNNNNNNNNN</td><td>1</td></tr>
<tr><td>Z</td><td>3920</td><td>hCoV-19/Wales/PHWC-4A2DD8/2020|EPI_ISL_744018|2020</td><td>····················</td><td>····················</td><td>····················</td><td>····················</td><td>NNNN················</td><td>NNNNNNNNNNNNNNNN····</td><td>1</td></tr>
<tr><td>Z</td><td>3922</td><td>hCoV-19/Wales/PHWC-4A2DF6/2020|EPI_ISL_744020|2020</td><td>····················</td><td>····················</td><td>····················</td><td>····················</td><td>·······NNNNNN·······</td><td>NNNNNNNNNNNNNNNN···N</td><td>1</td></tr>
<tr><td>Z</td><td>3924</td><td>hCoV-19/Wales/PHWC-4A2E11/2020|EPI_ISL_744022|2020</td><td>····················</td><td>····················</td><td>····················</td><td>····················</td><td>····················</td><td>NNNNN···············</td><td>1</td></tr>
<tr><td>Z</td><td>3925</td><td>hCoV-19/Wales/PHWC-4A2E20/2020|EPI_ISL_744023|2020</td><td>····················</td><td>····················</td><td>····················</td><td>····················</td><td>NNNNNNNNNNNNNNNNNNNN</td><td>NNNNNNNNNNNNNNNNNNNN</td><td>1</td></tr>
<tr><td>Z</td><td>3941</td><td>hCoV-19/Wales/PHWC-4A31F9/2020|EPI_ISL_744069|2020</td><td>····················</td><td>····················</td><td>····················</td><td>····················</td><td>NNNNNNNNNNNNNNNNNNNN</td><td>NNNNNNNNNNNNNNNNNNNN</td><td>1</td></tr>
<tr><td>Z</td><td>4099</td><td>hCoV-19/USA/CA-CZB-15288/2020|EPI_ISL_739502|2020-</td><td>····················</td><td>·A··················</td><td>····················</td><td>····················</td><td>NNNN················</td><td>····················</td><td>1</td></tr>
<tr><td>Z</td><td>4112</td><td>hCoV-19/USA/CA-CZB-13843/2020|EPI_ISL_739515|2020-</td><td>····················</td><td>····················</td><td>····················</td><td>····················</td><td>NNNN················</td><td>····················</td><td>1</td></tr>
<tr><td>Z</td><td>4118</td><td>hCoV-19/USA/CA-CZB-14999/2020|EPI_ISL_739521|2020-</td><td>····················</td><td>····················</td><td>····················</td><td>····················</td><td>NNNNNNNNNNNNNNNNNNNN</td><td>NNNNNNNNNNNNNNNNNNNN</td><td>1</td></tr>
<tr><td>Z</td><td>4119</td><td>hCoV-19/USA/CA-CZB-14405/2020|EPI_ISL_739522|2020-</td><td>····················</td><td>····················</td><td>····················</td><td>····················</td><td>NNNNNNNNNNNNNNNNNNNN</td><td>NNNNNNNNNNNNNNNNNNNN</td><td>1</td></tr>
<tr><td>Z</td><td>4120</td><td>hCoV-19/USA/CA-CZB-13792/2020|EPI_ISL_739523|2020-</td><td>····················</td><td>····················</td><td>····················</td><td>····················</td><td>NNNNNNNNNNNNNNNNNNNN</td><td>NNNNNNNNNNNNNNNN····</td><td>1</td></tr>
<tr><td>Z</td><td>4125</td><td>hCoV-19/USA/CA-CZB-13940/2020|EPI_ISL_739528|2020-</td><td>····················</td><td>····················</td><td>····················</td><td>····················</td><td>NNNNNNN·············</td><td>····················</td><td>1</td></tr>
<tr><td>Z</td><td>4126</td><td>hCoV-19/USA/CA-CZB-14897/2020|EPI_ISL_739529|2020-</td><td>····················</td><td>····················</td><td>····················</td><td>····················</td><td>NNNNNNNNNNNNNNNNN···</td><td>····················</td><td>1</td></tr>
<tr><td>Z</td><td>4142</td><td>hCoV-19/USA/CA-CZB-14229/2020|EPI_ISL_739545|2020-</td><td>····················</td><td>·A··················</td><td>····················</td><td>····················</td><td>NNNNNNNNNNNNNNNNN···</td><td>····················</td><td>1</td></tr>
<tr><td>Z</td><td>4152</td><td>hCoV-19/USA/CA-CZB-13836/2020|EPI_ISL_739555|2020-</td><td>····················</td><td>····················</td><td>····················</td><td>····················</td><td>NNNNNNNNNNNNNNNNNNNN</td><td>NNNNNNNNNNNNNNNNNNNN</td><td>1</td></tr>
<tr><td>Z</td><td>4157</td><td>hCoV-19/USA/CA-CZB-14053/2020|EPI_ISL_739590|2020-</td><td>····················</td><td>····················</td><td>····················</td><td>····················</td><td>NNNNNNNNNNNNNNNNNNNN</td><td>NNNNNNNNNNNNNNNNNNNN</td><td>1</td></tr>
<tr><td>Z</td><td>4284</td><td>hCoV-19/Wales/PHWC-4A2EC6/2020|EPI_ISL_744031|2020</td><td>····················</td><td>····················</td><td>····················</td><td>····················</td><td>NNNNNNNNNNNNNNNNNNNN</td><td>NNNNNNNNNNNNNNNNNNNN</td><td>1</td></tr>
<tr><td>Z</td><td>4289</td><td>hCoV-19/Wales/PHWC-4A2FC3/2020|EPI_ISL_744039|2020</td><td>····················</td><td>····················</td><td>····················</td><td>····················</td><td>NNNN················</td><td>NNNNNNNNNNNNNNN··NNN</td><td>1</td></tr>
<tr><td>Z</td><td>4290</td><td>hCoV-19/Wales/PHWC-4A2FD2/2020|EPI_ISL_744040|2020</td><td>····················</td><td>····················</td><td>····················</td><td>····················</td><td>NNNNNNNNNNNN········</td><td>NNNNNNNNNNNNNNNN····</td><td>1</td></tr>
<tr><td>Z</td><td>4310</td><td>hCoV-19/Wales/PHWC-4A32C9/2020|EPI_ISL_744082|2020</td><td>····················</td><td>····················</td><td>····················</td><td>····················</td><td>NNNN················</td><td>····················</td><td>1</td></tr>
<tr><td>Z</td><td>4313</td><td>hCoV-19/Wales/PHWC-4A3302/2020|EPI_ISL_744086|2020</td><td>····················</td><td>····················</td><td>····················</td><td>····················</td><td>NNNNNNNNNNNNNNNNNNNN</td><td>····················</td><td>1</td></tr>
<tr><td>Z</td><td>4316</td><td>hCoV-19/Wales/PHWC-4A3399/2020|EPI_ISL_744092|2020</td><td>····················</td><td>····················</td><td>····················</td><td>····················</td><td>NNNNNNNNNN···NNNNNNN</td><td>NNNNNNNNNN··NNNN····</td><td>1</td></tr>
<tr><td>Z</td><td>4318</td><td>hCoV-19/Wales/PHWC-4A33B7/2020|EPI_ISL_744094|2020</td><td>····················</td><td>····················</td><td>····················</td><td>····················</td><td>NNNN················</td><td>····················</td><td>1</td></tr>
<tr><td>Z</td><td>4360</td><td>hCoV-19/USA/CA-CZB-14162/2020|EPI_ISL_739567|2020-</td><td>····················</td><td>····················</td><td>····················</td><td>····················</td><td>NNNNNNNNNNNNNNNNNNNN</td><td>NNNNNNNNNNNNNNNNNNNN</td><td>1</td></tr>
<tr><td>Z</td><td>4368</td><td>hCoV-19/USA/CA-CZB-14266/2020|EPI_ISL_739575|2020-</td><td>····················</td><td>·A··················</td><td>····················</td><td>····················</td><td>NNNNNNNNNNNNNNNNN···</td><td>····················</td><td>1</td></tr>
<tr><td>Z</td><td>4369</td><td>hCoV-19/USA/CA-CZB-14130/2020|EPI_ISL_739576|2020-</td><td>····················</td><td>····················</td><td>····················</td><td>····················</td><td>NNNNNNNNNNNNNNNNNNNN</td><td>NNNNNNNNNNNNNNNN····</td><td>1</td></tr>
<tr><td>Z</td><td>4371</td><td>hCoV-19/USA/CA-CZB-14413/2020|EPI_ISL_739578|2020-</td><td>····················</td><td>····················</td><td>····················</td><td>····················</td><td>NNNNNNNNNNNNNNNNNNNN</td><td>NNNNNNNNNNNNNNNNNNNN</td><td>1</td></tr>
<tr><td>Z</td><td>4376</td><td>hCoV-19/USA/CA-CZB-13743/2020|EPI_ISL_739583|2020-</td><td>····················</td><td>·A··················</td><td>····················</td><td>····················</td><td>NNNNNNNNNNNNNN······</td><td>····················</td><td>1</td></tr>
<tr><td>Z</td><td>4382</td><td>hCoV-19/USA/CA-CZB-15323/2020|EPI_ISL_739589|2020-</td><td>····················</td><td>·A··················</td><td>····················</td><td>····················</td><td>NNNNNNNNNNNNNNNN····</td><td>NNNNNNNNNNNNNNNNNNNN</td><td>1</td></tr>
<tr><td>Z</td><td>4454</td><td>hCoV-19/Wales/PHWC-4A3487/2020|EPI_ISL_744102|2020</td><td>····················</td><td>····················</td><td>····················</td><td>····················</td><td>NNNNNNNNNNNNNNNNNNNN</td><td>NNNNN···············</td><td>1</td></tr>
<tr><td>Z</td><td>4456</td><td>hCoV-19/Wales/PHWC-4A350C/2020|EPI_ISL_744105|2020</td><td>····················</td><td>····················</td><td>····················</td><td>····················</td><td>NNNNNNNNNNNNNNNNNNNN</td><td>·················NNN</td><td>1</td></tr>
<tr><td>Z</td><td>4458</td><td>hCoV-19/Wales/PHWC-4A352A/2020|EPI_ISL_744107|2020</td><td>····················</td><td>····················</td><td>····················</td><td>····················</td><td>NNNNNNNNNNNNNNNNNNNN</td><td>····················</td><td>1</td></tr>
<tr><td>Z</td><td>4609</td><td>hCoV-19/USA/CA-CZB-14366/2020|EPI_ISL_739600|2020-</td><td>····················</td><td>·A··················</td><td>····················</td><td>····················</td><td>NNNNNNNNNNNNNNNNNNNN</td><td>NNNNNNNNNNNNNNNNNNNN</td><td>1</td></tr>
<tr><td>Z</td><td>4618</td><td>hCoV-19/USA/CA-CZB-15438/2020|EPI_ISL_739609|2020-</td><td>····················</td><td>·A··················</td><td>····················</td><td>····················</td><td>NNNNNNNNNNNNNNNNNNNN</td><td>NNNNNNNNNNNNNNNNNNNN</td><td>1</td></tr>
<tr><td>Z</td><td>4625</td><td>hCoV-19/USA/CA-CZB-13878/2020|EPI_ISL_739616|2020-</td><td>····················</td><td>·A··················</td><td>····················</td><td>····················</td><td>NNNNNNNNNNNNNNNNNNNN</td><td>NNNNNNNNNNNNNNNNNNNN</td><td>1</td></tr>
<tr><td>Z</td><td>4628</td><td>hCoV-19/USA/CA-CZB-14066/2020|EPI_ISL_739619|2020-</td><td>····················</td><td>····················</td><td>····················</td><td>····················</td><td>NNN·················</td><td>NNNNNNNNNNNNNNNNNNNN</td><td>1</td></tr>
<tr><td>Z</td><td>4632</td><td>hCoV-19/USA/CA-CZB-13744/2020|EPI_ISL_739623|2020-</td><td>····················</td><td>····················</td><td>····················</td><td>····················</td><td>NNNNNNNNNNNNNNNNNNNN</td><td>····················</td><td>1</td></tr>
<tr><td>Z</td><td>4640</td><td>hCoV-19/USA/CA-CZB-13871/2020|EPI_ISL_739631|2020-</td><td>····················</td><td>·A··················</td><td>····················</td><td>····················</td><td>NNNNNNNNNNNNNNNNNNNN</td><td>NNNNNNNNNNNNNNNNNNNN</td><td>1</td></tr>
<tr><td>Z</td><td>4655</td><td>hCoV-19/USA/CA-CZB-15301/2020|EPI_ISL_739647|2020-</td><td>····················</td><td>·A··················</td><td>····················</td><td>····················</td><td>NNNNNNNNNNNNNNNNNNNN</td><td>NNNNNNNNNNNNNNNNNNNN</td><td>1</td></tr>
<tr><td>Z</td><td>4665</td><td>hCoV-19/USA/CA-CZB-14574/2020|EPI_ISL_739657|2020-</td><td>····················</td><td>····················</td><td>····················</td><td>····················</td><td>NNNN················</td><td>NNNNNNNNNNNNNNNN····</td><td>1</td></tr>
<tr><td>Z</td><td>4667</td><td>hCoV-19/Colombia/DC-INS-4283/2020|EPI_ISL_739671|2</td><td>····················</td><td>····················</td><td>····················</td><td>····················</td><td>NNNNNNNNNNNNNNNNNNNN</td><td>NNNNNNNNNNNNNNNNNNNN</td><td>1</td></tr>
<tr><td>Z</td><td>4668</td><td>hCoV-19/Colombia/CAQ-INS-003003/2020|EPI_ISL_73967</td><td>····················</td><td>····················</td><td>····················</td><td>····················</td><td>NNNNNNNNNNNNNNNNNNNN</td><td>NNNNNNNNNNNNNNNNNNNN</td><td>1</td></tr>
<tr><td>Z</td><td>4669</td><td>hCoV-19/Colombia/DC-INS-4301/2020|EPI_ISL_739673|2</td><td>····················</td><td>·A··················</td><td>····················</td><td>····················</td><td>NNNNNNNNNNNNNNNNNNNN</td><td>NNNNNNNNNNNNNNNNNNNN</td><td>1</td></tr>
<tr><td>Z</td><td>4940</td><td>hCoV-19/Wales/PHWC-49B8C2/2020|EPI_ISL_742400|2020</td><td>····················</td><td>····················</td><td>····················</td><td>····················</td><td>NNNNNNNNNNNNNNNNNNNN</td><td>NNNNNNNNNNNNNNNNNNNN</td><td>1</td></tr>
<tr><td>Z</td><td>4942</td><td>hCoV-19/Wales/PHWC-49B8E0/2020|EPI_ISL_742402|2020</td><td>····················</td><td>····················</td><td>····················</td><td>····················</td><td>NNNNNNNNNNNNNNNNNNNN</td><td>NNNNNNNNNNNNNNNNNNNN</td><td>1</td></tr>
<tr><td>Z</td><td>4944</td><td>hCoV-19/Wales/PHWC-49B956/2020|EPI_ISL_742406|2020</td><td>····················</td><td>····················</td><td>····················</td><td>····················</td><td>NNNNNNNNNNNNNNNNNNNN</td><td>NNNNNNNNNNNNNNNNNNNN</td><td>1</td></tr>
<tr><td>Z</td><td>4947</td><td>hCoV-19/Wales/PHWC-49B9A1/2020|EPI_ISL_742410|2020</td><td>····················</td><td>····················</td><td>····················</td><td>····················</td><td>NNNNNNNNNNNNNNNNNNNN</td><td>NNNNNNNNNNNNNNNNNNNN</td><td>1</td></tr>
<tr><td>Z</td><td>4948</td><td>hCoV-19/Wales/PHWC-49B9B0/2020|EPI_ISL_742411|2020</td><td>····················</td><td>····················</td><td>····················</td><td>····················</td><td>NNNNNNNNNNNNNNNNNNNN</td><td>NNNNNNNNNNNNNNNNNNNN</td><td>1</td></tr>
<tr><td>Z</td><td>4949</td><td>hCoV-19/Wales/PHWC-49B9CF/2020|EPI_ISL_742412|2020</td><td>····················</td><td>····················</td><td>····················</td><td>····················</td><td>NNNNNNNNNNNNNNNNNNNN</td><td>NNNNNNNNNNNNNNNNNNNN</td><td>1</td></tr>
<tr><td>Z</td><td>4951</td><td>hCoV-19/Wales/PHWC-49B9FC/2020|EPI_ISL_742414|2020</td><td>····················</td><td>····················</td><td>····················</td><td>····················</td><td>NNNNNNNNNNNNNNNNNNNN</td><td>NNNNNNNNNNNNNNNNNNNN</td><td>1</td></tr>
<tr><td>Z</td><td>4952</td><td>hCoV-19/Wales/PHWC-49BA17/2020|EPI_ISL_742416|2020</td><td>····················</td><td>····················</td><td>····················</td><td>····················</td><td>NNNNNNNNNNNNNNNNNNNN</td><td>NNNNNNNNNNNNNNNNNNNN</td><td>1</td></tr>
<tr><td>Z</td><td>4954</td><td>hCoV-19/Wales/PHWC-49BA35/2020|EPI_ISL_742418|2020</td><td>····················</td><td>····················</td><td>····················</td><td>····················</td><td>NNNNNNNNNNNNNNNNNNNN</td><td>NNNNNNNNNN··········</td><td>1</td></tr>
<tr><td>Z</td><td>4956</td><td>hCoV-19/Wales/PHWC-49BA62/2020|EPI_ISL_742421|2020</td><td>····················</td><td>····················</td><td>····················</td><td>····················</td><td>NNNNNNNNNNNNNNNNNNN·</td><td>NNNNNNNNNNNNNNNNNNNN</td><td>1</td></tr>
<tr><td>Z</td><td>4957</td><td>hCoV-19/Wales/PHWC-49BA71/2020|EPI_ISL_742422|2020</td><td>····················</td><td>····················</td><td>····················</td><td>····················</td><td>NNNNNNNNNNNNNNNNNNNN</td><td>NNNNNNNNNNNNNNNNNNNN</td><td>1</td></tr>
<tr><td>Z</td><td>4960</td><td>hCoV-19/Wales/PHWC-49BAAE/2020|EPI_ISL_742425|2020</td><td>····················</td><td>····················</td><td>····················</td><td>····················</td><td>NNNNNNNNNNNNNNNNNNNN</td><td>NNNNNNNNNNNNNNNNNNNN</td><td>1</td></tr>
<tr><td>Z</td><td>4963</td><td>hCoV-19/Wales/PHWC-49BAEA/2020|EPI_ISL_742429|2020</td><td>····················</td><td>····················</td><td>····················</td><td>····················</td><td>NNNNNNNNNNNNNNNNNNNN</td><td>NNNNNNNNNNNNNNNNNNNN</td><td>1</td></tr>
<tr><td>Z</td><td>4964</td><td>hCoV-19/Wales/PHWC-49BAF9/2020|EPI_ISL_742430|2020</td><td>····················</td><td>····················</td><td>····················</td><td>····················</td><td>NNNNNNNNNNNNNNNNNNNN</td><td>NNNNNNNNNNNNNNNNNNNN</td><td>1</td></tr>
<tr><td>Z</td><td>4967</td><td>hCoV-19/Wales/PHWC-49BB32/2020|EPI_ISL_742434|2020</td><td>····················</td><td>····················</td><td>····················</td><td>····················</td><td>····NNNNNNNNNNNNNNNN</td><td>NNNNNNNNNNN··NNNNNNN</td><td>1</td></tr>
<tr><td>Z</td><td>4970</td><td>hCoV-19/Wales/PHWC-49BB7E/2020|EPI_ISL_742438|2020</td><td>····················</td><td>····················</td><td>····················</td><td>····················</td><td>NNNNNNNNNNNNNNNNNNNN</td><td>NNNNNNNNNNN········N</td><td>1</td></tr>
<tr><td>Z</td><td>4972</td><td>hCoV-19/Wales/PHWC-49BB9C/2020|EPI_ISL_742440|2020</td><td>····················</td><td>····················</td><td>····················</td><td>····················</td><td>NNNNNNNNNNNNNNNNNNNN</td><td>NNNNNNNNNNNNNNNNNNNN</td><td>1</td></tr>
<tr><td>Z</td><td>4974</td><td>hCoV-19/Wales/PHWC-49BBD8/2020|EPI_ISL_742443|2020</td><td>····················</td><td>····················</td><td>····················</td><td>····················</td><td>NNNNNNNNNNNNNNNNNNNN</td><td>NNNNNNNNNNNNNNNNNNNN</td><td>1</td></tr>
<tr><td>Z</td><td>4975</td><td>hCoV-19/Wales/PHWC-49BBF6/2020|EPI_ISL_742444|2020</td><td>····················</td><td>····················</td><td>····················</td><td>····················</td><td>NNNNNNNNNNNNNNNNNNNN</td><td>NNNNNNNNNNNNNNNNNNNN</td><td>1</td></tr>
<tr><td>Z</td><td>4976</td><td>hCoV-19/Wales/PHWC-49BC02/2020|EPI_ISL_742445|2020</td><td>····················</td><td>····················</td><td>····················</td><td>····················</td><td>NNNNNNNNNNNNNNNNNNNN</td><td>·····NNNNNNNNNNNNNNN</td><td>1</td></tr>
<tr><td>Z</td><td>4980</td><td>hCoV-19/Wales/PHWC-49BC4E/2020|EPI_ISL_742449|2020</td><td>····················</td><td>····················</td><td>····················</td><td>····················</td><td>NNNNNNNNNNNNNNNNNNNN</td><td>NNNNNNNNNNNNNNNNNNNN</td><td>1</td></tr>
<tr><td>Z</td><td>4983</td><td>hCoV-19/Wales/PHWC-49BC7B/2020|EPI_ISL_742452|2020</td><td>····················</td><td>····················</td><td>····················</td><td>····················</td><td>NNNNNNNNNNNNNNNNNNNN</td><td>NNNNNNNNNNNNNNNNNNNN</td><td>1</td></tr>
<tr><td>Z</td><td>4985</td><td>hCoV-19/Wales/PHWC-49BC99/2020|EPI_ISL_742454|2020</td><td>····················</td><td>····················</td><td>····················</td><td>····················</td><td>NNNNNNNNNNNNNNNNNNNN</td><td>NNNNNNNNNNNNNNN·NNNN</td><td>1</td></tr>
<tr><td>Z</td><td>4988</td><td>hCoV-19/Wales/PHWC-49BCF3/2020|EPI_ISL_742458|2020</td><td>····················</td><td>····················</td><td>····················</td><td>····················</td><td>NNNNNNNNNNNNNNNNNNNN</td><td>NNNNNNNNNNNNNNNNNNNN</td><td>1</td></tr>
<tr><td>Z</td><td>4992</td><td>hCoV-19/Wales/PHWC-49BD87/2020|EPI_ISL_742465|2020</td><td>····················</td><td>····················</td><td>····················</td><td>····················</td><td>NNNNNNNNNNNNNNNNNNNN</td><td>NNNNNNNNNNNNNNNNNNNN</td><td>1</td></tr>
<tr><td>Z</td><td>4993</td><td>hCoV-19/Wales/PHWC-49BDC3/2020|EPI_ISL_742466|2020</td><td>·T··················</td><td>····················</td><td>····················</td><td>····················</td><td>NNNNNNNNNNNNNNNNNNNN</td><td>NNNNNNNNNNNNNNNNNNNN</td><td>1</td></tr>
<tr><td>Z</td><td>4996</td><td>hCoV-19/Wales/PHWC-49BDF0/2020|EPI_ISL_742469|2020</td><td>····················</td><td>····················</td><td>····················</td><td>····················</td><td>NNNNNNNNNNNNNNNNNNNN</td><td>NNNNNNNNNNNNNNNNNNNN</td><td>1</td></tr>
<tr><td>Z</td><td>4997</td><td>hCoV-19/Wales/PHWC-49BE0C/2020|EPI_ISL_742470|2020</td><td>····················</td><td>····················</td><td>····················</td><td>····················</td><td>NNNNNNNNNNNNNNNNNNNN</td><td>NNNNNNN·············</td><td>1</td></tr>
<tr><td>Z</td><td>5000</td><td>hCoV-19/Wales/PHWC-49BE57/2020|EPI_ISL_742473|2020</td><td>····················</td><td>····················</td><td>····················</td><td>····················</td><td>NNNNNNNNNNNNNNNNNNNN</td><td>NNNNNNNNNNNNNNNNNNNN</td><td>1</td></tr>
<tr><td>Z</td><td>5002</td><td>hCoV-19/Wales/PHWC-49BE75/2020|EPI_ISL_742475|2020</td><td>····················</td><td>····················</td><td>····················</td><td>····················</td><td>NNNNNNNNNNNNNNNNNNNN</td><td>NNNNNNNNNNNNNNNNNNNN</td><td>1</td></tr>
<tr><td>Z</td><td>5006</td><td>hCoV-19/Wales/PHWC-49BEEE/2020|EPI_ISL_742480|2020</td><td>····················</td><td>····················</td><td>····················</td><td>····················</td><td>····················</td><td>NNNNNNNNNNNNNNNN····</td><td>1</td></tr>
<tr><td>Z</td><td>5008</td><td>hCoV-19/Wales/PHWC-49BF09/2020|EPI_ISL_742482|2020</td><td>····················</td><td>····················</td><td>····················</td><td>····················</td><td>NNNNNNNNNNNNNNNNNNNN</td><td>NNNNNNNNNNNNNNNNNNNN</td><td>1</td></tr>
<tr><td>Z</td><td>5011</td><td>hCoV-19/Wales/PHWC-49BF54/2020|EPI_ISL_742485|2020</td><td>····················</td><td>····················</td><td>····················</td><td>····················</td><td>NNNNNNNNNNNNNNNNNNNN</td><td>NNNNNNNNNNNNNNNNNNNN</td><td>1</td></tr>
<tr><td>Z</td><td>5013</td><td>hCoV-19/Wales/PHWC-49BF81/2020|EPI_ISL_742487|2020</td><td>····················</td><td>····················</td><td>····················</td><td>····················</td><td>NNNNNNNNNNNNNNNNNNNN</td><td>NNNNNNNNNNNNNNNNNNNN</td><td>1</td></tr>
</table>

| | | | | | | | | |
|---|---|---|---|---|---|---|---|---|
| Z | 5015 hCoV-19/Wales/PHWC-49BFAF/2020\|EPI_ISL_742489\|2020 | ···················· | ···················· | ···················· | ···················· | NNNNNNNNNNNNNNNNNNNN | ···················· | 1 |
| Z | 5016 hCoV-19/Wales/PHWC-49BFBE/2020\|EPI_ISL_742490\|2020 | ···················· | ···················· | ···················· | ···················· | NNNNNNNNNNNNNNNNNNNN | NNNNNNNNNNNNNNNNNNNN | 1 |
| Z | 5019 hCoV-19/Wales/PHWC-49BFFA/2020\|EPI_ISL_742494\|2020 | ···················· | ···················· | ···················· | ···················· | NNNNNNNNNNNNNNNNNNNN | NNNNNNNNNNNNNNNNNNNN | 1 |
| Z | 5023 hCoV-19/Wales/PHWC-49C041/2020\|EPI_ISL_742498\|2020 | ···················· | ···················· | ···················· | ···················· | NNNNNNNNNNNNNNNNNNNN | NNNNNNNNNNNNNNNNNNNN | 1 |
| Z | 5024 hCoV-19/Wales/PHWC-49C050/2020\|EPI_ISL_742499\|2020 | ···················· | ···················· | ···················· | ···················· | ···················· | NNNNNN···NNNNNN····· | 1 |
| Z | 5027 hCoV-19/Wales/PHWC-4A1629/2020\|EPI_ISL_743702\|2020 | ···················· | ···················· | ···················· | ···················· | NNNNNNNNNNNNNNNNNNNN | NNNNNNNNNNNNNNNNN··· | 1 |
| Z | 5034 hCoV-19/Wales/PHWC-4A1692/2020\|EPI_ISL_743709\|2020 | ···················· | ···················· | ···················· | ···················· | NNNNNNNNNNNNNNNNNNNN | ···················· | 1 |
| Z | 5046 hCoV-19/Wales/PHWC-4A17EA/2020\|EPI_ISL_743728\|2020 | ···················· | ···················· | ···················· | ···················· | NNNNNNNNNNNNNNNNNNNN | NNNNNNNNNNNNNNNNNNNN | 1 |
| Z | 5047 hCoV-19/Wales/PHWC-4A17F9/2020\|EPI_ISL_743729\|2020 | ···················· | ···················· | ···················· | ···················· | NNNNNNNNNNNNNNNNNNNN | NNNNNNNNNNNNNNNNNNNN | 1 |
| Z | 5056 hCoV-19/Wales/PHWC-4A189C/2020\|EPI_ISL_743738\|2020 | ···················· | ···················· | ···················· | ···················· | ···················· | NNNNNNNNNNNNNNN····· | 1 |
| Z | 5065 hCoV-19/Wales/PHWC-4A198A/2020\|EPI_ISL_743753\|2020 | ···················· | ···················· | ···················· | ···················· | NNNNNNNNNNNNNNNNNNNN | NNNNNNNNNNNNNNNNNNNN | 1 |
| Z | 5070 hCoV-19/Wales/PHWC-4A19D5/2020\|EPI_ISL_743758\|2020 | ···················· | ···················· | ···················· | ···················· | NNNNNNNNNNNNNNNNNNNN | ···················· | 1 |
| Z | 5081 hCoV-19/Wales/PHWC-4A1AB4/2020\|EPI_ISL_743772\|2020 | ···················· | ···················· | ···················· | ···················· | ···················· | NNNNN··············· | 1 |
| Z | 5085 hCoV-19/Wales/PHWC-4A1B1B/2020\|EPI_ISL_743778\|2020 | ···················· | ···················· | ···················· | ···················· | NNNNNNNNNNNNNNNNNNNN | NNNNNNNNNNNNNNNNNNNN | 1 |
| Z | 5090 hCoV-19/Wales/PHWC-4A1B66/2020\|EPI_ISL_743783\|2020 | ···················· | ···················· | ···················· | ···················· | NNNNNNNNNNNNNNNNNNNN | NNNNNNNNNNNNNNNNNNNN | 1 |
| Z | 5093 hCoV-19/Wales/PHWC-4A1BC0/2020\|EPI_ISL_743787\|2020 | ···················· | ···················· | ···················· | ···················· | NNNNNNNNNNNNNNNNNNNN | NNNNNNNNNNNNNNNNNNNN | 1 |
| Z | 5096 hCoV-19/Wales/PHWC-4A1C09/2020\|EPI_ISL_743790\|2020 | ···················· | ···················· | ···················· | ···················· | ··NNNNNNNNNNNNNNNNNN | NNNNNNNNNNNNNNN····· | 1 |
| Z | 5101 hCoV-19/Wales/PHWC-4A1C72/2020\|EPI_ISL_743797\|2020 | ···················· | ···················· | ···················· | ···················· | NNNNNNNNNNNNNNNNNNNN | NNNNNNNNNNNNNNNNNNNN | 1 |
| Z | 5102 hCoV-19/Wales/PHWC-4A1C81/2020\|EPI_ISL_743798\|2020 | ···················· | ···················· | ···················· | ···················· | NNNNNNNNNNNNNNNNNNNN | NNNNNNNNNNNNNNNNNNNN | 1 |
| Z | 5144 hCoV-19/Wales/PHWC-49C08D/2020\|EPI_ISL_742502\|2020 | ···················· | ···················· | ···················· | ···················· | NNNNNNN············· | ···················· | 1 |
| Z | 5146 hCoV-19/Wales/PHWC-49C0BA/2020\|EPI_ISL_742505\|2020 | ···················· | ···················· | ···················· | ···················· | NNNNNNNNNNNNNNNNNNNN | NNNNNNNNNNNNNNNNNNNN | 1 |
| Z | 5147 hCoV-19/Wales/PHWC-49C0C9/2020\|EPI_ISL_742506\|2020 | ···················· | ···················· | ···················· | ···················· | NNNNNNNNNNNNNNNNNNNN | NNNNNNNNNNNNNNNNNNNN | 1 |
| Z | 5148 hCoV-19/Wales/PHWC-49C0E7/2020\|EPI_ISL_742507\|2020 | ···················· | ···················· | ···················· | ···················· | NNNNNNNNNNNNNNNNNNNN | NNNNNNNNNNNNNNNNNNNN | 1 |
| Z | 5150 hCoV-19/Wales/PHWC-49C111/2020\|EPI_ISL_742510\|2020 | ···················· | ···················· | ···················· | ···················· | NNNNNNNNNNNNNNNNNNNN | NNNNNNNNNNNNNNNNNNNN | 1 |
| Z | 5151 hCoV-19/Wales/PHWC-49C120/2020\|EPI_ISL_742511\|2020 | ···················· | ···················· | ···················· | ···················· | NNNNNNNNNNNNNNNNNNNN | NNNNNNNNNNNNNNNNNNNN | 1 |
| Z | 5152 hCoV-19/Wales/PHWC-49C13F/2020\|EPI_ISL_742512\|2020 | ···················· | ···················· | ···················· | ···················· | NNNNNNNNNNNNNNNNNNNN | NNNNNNNNNNNNNNNNNNNN | 1 |
| Z | 5155 hCoV-19/Wales/PHWC-49C18A/2020\|EPI_ISL_742517\|2020 | ···················· | ···················· | ···················· | ···················· | NNNNNNNNNNNNNNNNNNNN | NNNNNNNNNNNNNNNNNNNN | 1 |
| Z | 5156 hCoV-19/Wales/PHWC-49C199/2020\|EPI_ISL_742518\|2020 | ···················· | ···················· | ···················· | ···················· | NNNNNNNNNNNNNNNNNNNN | NNNNNNNNNNNNNNNNNNNN | 1 |
| Z | 5160 hCoV-19/Wales/PHWC-49C1D5/2020\|EPI_ISL_742522\|2020 | ···················· | ···················· | ···················· | ···················· | NNNNNNNNNNNNNNNNNNNN | NNNNNNNNNNNNNNNNNNNN | 1 |
| Z | 5163 hCoV-19/Wales/PHWC-49C22D/2020\|EPI_ISL_742527\|2020 | ···················· | ···················· | ···················· | ···················· | NNNNNNNNNNNNNNNNNNNN | NNNNNNNNNNNNNNNNNNNN | 1 |
| Z | 5164 hCoV-19/Wales/PHWC-49C23C/2020\|EPI_ISL_742528\|2020 | ···················· | ···················· | ···················· | ···················· | NNNNNNNNNNNN········ | ···················· | 1 |
| Z | 5165 hCoV-19/Wales/PHWC-49C269/2020\|EPI_ISL_742531\|2020 | ···················· | ···················· | ···················· | ···················· | NNNNNNNNNNNNNNNNNNNN | NNNNNNNNNNNNNNNNNNNN | 1 |
| Z | 5167 hCoV-19/Wales/PHWC-49C296/2020\|EPI_ISL_742534\|2020 | ···················· | ···················· | ···················· | ···················· | NNNNNNNNNNNNNNNNNNNN | NNNNNNNNNNNNNNNNNNNN | 1 |
| Z | 5169 hCoV-19/Wales/PHWC-49C2C3/2020\|EPI_ISL_742537\|2020 | ···················· | ···················· | ···················· | ···················· | NNNNNNNNNNNNNNNNNNNN | NNNNNNNNNNNNNNNNNNNN | 1 |
| Z | 5171 hCoV-19/Wales/PHWC-49C2F0/2020\|EPI_ISL_742540\|2020 | ···················· | ···················· | ···················· | ···················· | NNNNNNNNNNNNNNNNNNNN | NNNNNNNNNNNNNNNNNNNN | 1 |
| Z | 5192 hCoV-19/Wales/PHWC-49C4DC/2020\|EPI_ISL_742563\|2020 | ···················· | ···················· | ···················· | ···················· | ···················· | NNNNNNNNNNNNNNN····· | 1 |
| Z | 5209 hCoV-19/Wales/PHWC-49C65E/2020\|EPI_ISL_742586\|2020 | ···················· | ···················· | ···················· | ···················· | N······NNNNNNNNNNNNN | NNNNNNNNNNNNNNNNNNNN | 1 |
| Z | 5221 hCoV-19/Wales/PHWC-4A1CFA/2020\|EPI_ISL_743804\|2020 | ···················· | ···················· | ···················· | ···················· | NNNNNNNNNNNNNNNNNNNN | ···················· | 1 |
| Z | 5223 hCoV-19/Wales/PHWC-4A1D42/2020\|EPI_ISL_743807\|2020 | ···················· | ···················· | ···················· | ···················· | NNNNNNNNNNNNNNNNNNNN | NNNNNNNNNNNNNNNNNNNN | 1 |
| Z | 5235 hCoV-19/Wales/PHWC-4A1E7C/2020\|EPI_ISL_743825\|2020 | ···················· | ···················· | ···················· | ···················· | NNNNNNNNNNNNNNNNNNNN | NNNNNNNNNNNNNNNNNNNN | 1 |
| Z | 5246 hCoV-19/Wales/PHWC-4A1FD3/2020\|EPI_ISL_743840\|2020 | ···················· | ···················· | ···················· | ···················· | NNNNNNNNNNNNNNNNNNNN | NNNNNNNNNNNNNNNNNNNN | 1 |
| Z | 5248 hCoV-19/Wales/PHWC-4A200C/2020\|EPI_ISL_743842\|2020 | ···················· | ···················· | ···················· | ···················· | NNNNNNNNNNNNNNNNNNNN | NNNNNNNNNNNNNNNNNNNN | 1 |
| Z | 5249 hCoV-19/Wales/PHWC-4A2084/2020\|EPI_ISL_743843\|2020 | ···················· | ···················· | ···················· | ···················· | ···················· | NNNNNNNNNNNNNNNNNNNN | 1 |
| Z | 5250 hCoV-19/Wales/PHWC-4A20EE/2020\|EPI_ISL_743846\|2020 | ···················· | ···················· | ···················· | ···················· | NNNNNNNNNNNNNNNNNNNN | NNNNNNNNNNNNNNNNNNNN | 1 |
| Z | 5251 hCoV-19/Wales/PHWC-4A2118/2020\|EPI_ISL_743848\|2020 | ···················· | ···················· | ···················· | ···················· | NNNNNNNNNNNNNNNNNNNN | NNNNNNNNNNNNNNNNNNNN | 1 |
| Z | 5252 hCoV-19/Wales/PHWC-4A2127/2020\|EPI_ISL_743849\|2020 | ···················· | ···················· | ···················· | ···················· | NNNNNNNNNNNNNNNNNNNN | NNNNNNNNNNNNNNNNNNNN | 1 |
| Z | 5253 hCoV-19/Wales/PHWC-4A2145/2020\|EPI_ISL_743850\|2020 | ···················· | ···················· | ···················· | ···················· | ···················· | NNNNNNNNNNNNNNN····· | 1 |
| Z | 5254 hCoV-19/Wales/PHWC-4A2154/2020\|EPI_ISL_743851\|2020 | ···················· | ···················· | ···················· | ···················· | NNNNNNNNNNNNNNNNNNNN | NNNNNNNNNNNNNNNNNNNN | 1 |
| Z | 5263 hCoV-19/Wales/PHWC-4A2206/2020\|EPI_ISL_743862\|2020 | ···················· | ···················· | ···················· | ···················· | NNNNNNNNNNNNNNNNNNNN | NNNNNNNNNNNNNNNNNNNN | 1 |
| Z | 5267 hCoV-19/Wales/PHWC-4A229D/2020\|EPI_ISL_743867\|2020 | ···················· | ···················· | ···················· | ···················· | NNNNNNNNNNNNNNNNNNNN | NNNNNNNNNNNNNNNNNNNN | 1 |
| Z | 5279 hCoV-19/Wales/PHWC-4A23C7/2020\|EPI_ISL_743882\|2020 | ···················· | ···················· | ···················· | ···················· | NNNNNNNNNNNNNNNNNNNN | ···················· | 1 |
| Z | 5380 hCoV-19/USA/CA-CZB-14988/2020\|EPI_ISL_739031\|2020- | ···················· | ·A·················· | ···················· | ···················· | NNNNNNNNNNNNNNNNNNNN | NNNNNNNNNNNNNNNNNNNN | 1 |
| Z | 5384 hCoV-19/USA/CA-CZB-13999/2020\|EPI_ISL_739035\|2020- | ···················· | ···················· | ···················· | ···················· | NNNNNNNNNNNNNNNNNNNN | NNNNNNNNNNNNNNNNNNNN | 1 |
| Z | 5422 hCoV-19/Wales/PHWC-49C7A6/2020\|EPI_ISL_742601\|2020 | ···················· | ···················· | ···················· | ···················· | NNNNNNNNNNNNNNNNNNNN | ···················· | 1 |
| Z | 5423 hCoV-19/Wales/PHWC-49C7B5/2020\|EPI_ISL_742602\|2020 | ···················· | ···················· | ···················· | ···················· | NNNNNNNNNNNNNNNNNNNN | ···················· | 1 |
| Z | 5437 hCoV-19/Wales/PHWC-49C982/2020\|EPI_ISL_742623\|2020 | ···················· | ···················· | ···················· | ···················· | NNNNNNNNNNNNNNNNNNNN | ··········NNNNN····· | 1 |
| Z | 5442 hCoV-19/Wales/PHWC-49C9FB/2020\|EPI_ISL_742630\|2020 | ···················· | ···················· | ···················· | ···················· | NNNNNNNNNNNNNNNNNNNN | NNNNNNNNNNNNNNN····· | 1 |
| Z | 5445 hCoV-19/Wales/PHWC-49CA61/2020\|EPI_ISL_742637\|2020 | ···················· | ···················· | ···················· | ···················· | NNNNNNNNNNNNNNNNNNNN | ···················· | 1 |
| Z | 5448 hCoV-19/Wales/PHWC-49CA9E/2020\|EPI_ISL_742640\|2020 | ···················· | ···················· | ···················· | ···················· | NNNNNNNNNNNNNNNNNNNN | ···NNNNNNNNNNNN····· | 1 |
| Z | 5456 hCoV-19/Wales/PHWC-49CCC5/2020\|EPI_ISL_742673\|2020 | ···················· | ···················· | ···················· | ···················· | NNNNNNNNNNNNNNNNNNNN | NNNNNNNNNNNNNNNNNNNN | 1 |
| Z | 5458 hCoV-19/Wales/PHWC-49CD0E/2020\|EPI_ISL_742677\|2020 | ···················· | ···················· | ···················· | ···················· | NNNNNNNNNNNNNNNNNNNN | NNNNNNNNNNNNNNNNNNNN | 1 |
| Z | 5459 hCoV-19/Wales/PHWC-49CD1D/2020\|EPI_ISL_742678\|2020 | ···················· | ···················· | ···················· | ···················· | NNNNNNNNNNNNNNNNNNNN | ···················· | 1 |
| Z | 5461 hCoV-19/Wales/PHWC-49CD3B/2020\|EPI_ISL_742680\|2020 | ···················· | ···················· | ···················· | ···················· | ···················· | NNNNNNNNNNNNNNN····· | 1 |
| Z | 5465 hCoV-19/Wales/PHWC-49CD77/2020\|EPI_ISL_742684\|2020 | ···················· | ···················· | ···················· | ···················· | NNNN·N·············· | ···················· | 1 |
| Z | 5471 hCoV-19/Wales/PHWC-4A252B/2020\|EPI_ISL_743902\|2020 | ···················· | ···················· | ···················· | ···················· | NNNNNNNNNNNNNNNNNNNN | NNNNNNNNNNNNNNNNNNNN | 1 |
| Z | 5523 hCoV-19/Wales/PHWC-4A2A0E/2020\|EPI_ISL_743966\|2020 | ···················· | ···················· | ···················· | ···················· | NNNN·NNNN··········· | ···················· | 1 |
| Z | 5532 hCoV-19/Wales/PHWC-4A2C62/2020\|EPI_ISL_743999\|2020 | ···················· | ···················· | ···················· | ···················· | NNNNNNNNNNNNNNNNNNNN | ···················· | 1 |
| Z | 5637 hCoV-19/USA/CA-CZB-14593/2020\|EPI_ISL_739004\|2020- | ···················· | ···················· | ···················· | ···················· | NNNNNNNNNNNNNNNNNNNN | NNNNNNNNNNNNNNNNNNNN | 1 |
| Z | 5657 hCoV-19/USA/CA-CZB-15313/2020\|EPI_ISL_739025\|2020- | ···················· | ·A·················· | ···················· | ···················· | NNNNNNNNNNNNNNNNNNNN | ···················· | 1 |
| Z | 5658 hCoV-19/USA/CA-CZB-14957/2020\|EPI_ISL_739026\|2020- | ···················· | ···················· | ···················· | ···················· | NNNNNNNNNNNNNNNNNNNN | ·····NNNNNNNNNNNNNNN | 1 |
| Z | 5662 hCoV-19/USA/CA-CZB-15304/2020\|EPI_ISL_739041\|2020- | ···················· | ·A·················· | ···················· | ···················· | NNNNNNNNNNNNNNNNNNNN | NNNNNNN············· | 1 |
| Z | 5678 hCoV-19/USA/CA-CZB-14070/2020\|EPI_ISL_739057\|2020- | ···················· | ···················· | ···················· | ···················· | NNNNNNNNNNNNNNNNNNNN | ···················· | 1 |
| Z | 5683 hCoV-19/USA/CA-CZB-13764/2020\|EPI_ISL_739062\|2020- | ···················· | ···················· | ···················· | ···················· | NNNNNNNNNNNNNNNNNNNN | NNNNNNNNNNNNNNNNNNNN | 1 |
| Z | 5685 hCoV-19/USA/CA-CZB-13947/2020\|EPI_ISL_739064\|2020- | ···················· | ···················· | ···················· | ···················· | NNNNNNNNNNNNNNNNNNNN | NNNNNNNNNNNNNNNNNNNN | 1 |
| Z | 5686 hCoV-19/USA/CA-CZB-15188/2020\|EPI_ISL_739065\|2020- | ···················· | ···················· | ···················· | ···················· | NNNNNNNNNNNNNNNNNNNN | NNNNNNNNNNNNNNNNNNNN | 1 |
| Z | 5688 hCoV-19/USA/CA-CZB-15322/2020\|EPI_ISL_739067\|2020- | ···················· | ·A·················· | ···················· | ···················· | NNNNNNNNNNNNNNNNNNNN | NNNNNNNNNNNNNNNNNNNN | 1 |
| Z | 5709 hCoV-19/USA/CA-CZB-14963/2020\|EPI_ISL_739091\|2020- | ···················· | ·A·················· | ···················· | ···················· | NNNNNNNNNNNNNNNNNNNN | NNNNNNNNNNNNNNNNNNNN | 1 |
| Z | 5714 hCoV-19/USA/CA-CZB-13890/2020\|EPI_ISL_739096\|2020- | ···················· | ···················· | ···················· | ···················· | NNNNNNNNNNNNNNNNNNNN | NNNNNNNNNNNNNNNNNNNN | 1 |
| Z | 5846 hCoV-19/Wales/PHWC-49CB9B/2020\|EPI_ISL_742654\|2020 | ···················· | ···················· | ···················· | ···················· | NNNNNNNNNNNNNNNNNNNN | NNNNNNNNNNNNNNNNNNNN | 1 |
| Z | 5851 hCoV-19/Wales/PHWC-49CBE6/2020\|EPI_ISL_742659\|2020 | ···················· | ···················· | ···················· | ···················· | ···················· | NNNNNNNNNNN·····NNNN | 1 |

| | | | | | | | | |
|---|---|---|---|---|---|---|---|---|
| Z | 5866 hCoV-19/Wales/PHWC-49CE38/2020\|EPI_ISL_742695\|2020 | ···················· | ···················· | ···················· | ···················· | ···················· | ·········NNNNNN····· | 1 |
| Z | 5885 hCoV-19/Wales/PHWC-4A2C17/2020\|EPI_ISL_743995\|2020 | ···················· | ···················· | ···················· | ···················· | ···················· | NNNNN··············· | 1 |
| Z | 5891 hCoV-19/USA/CA-CZB-14881/2020\|EPI_ISL_739089\|2020- | ···················· | ···················· | ···················· | ···················· | NNNNNNNNNNNNNNNNNNNN | ···················· | 1 |
| Z | 5902 hCoV-19/USA/CA-CZB-13749/2020\|EPI_ISL_739110\|2020- | ···················· | ·A·················· | ···················· | ···················· | NNNNNNNNNNNNNNNNNNNN | NNNNNNNNNNNNNNNNNNNN | 1 |
| Z | 5905 hCoV-19/USA/CA-CZB-14300/2020\|EPI_ISL_739113\|2020- | ···················· | ·A·················· | ···················· | ···················· | NNNNNNNNNNNNNNNNNNNN | ···················· | 1 |
| Z | 5910 hCoV-19/USA/CA-CZB-13941/2020\|EPI_ISL_739118\|2020- | ···················· | ···················· | ···················· | ···················· | NNNNNNNNNNNNNNNNNNNN | ···················· | 1 |
| Z | 5916 hCoV-19/USA/CA-CZB-15243/2020\|EPI_ISL_739152\|2020- | ···················· | ···················· | ···················· | ···················· | NNNN················ | ···················· | 1 |
| Z | 5989 hCoV-19/Wales/PHWC-49CF17/2020\|EPI_ISL_742707\|2020 | ···················· | ···················· | ···················· | ···················· | NNNNNNNNNNNNNNNNNNNN | NNNNNNNNNNNNNNNNNNNN | 1 |
| Z | 6010 hCoV-19/Wales/PHWC-49D0C8/2020\|EPI_ISL_742731\|2020 | ···················· | ···················· | ···················· | ···················· | ···················· | ···········NNNN····· | 1 |
| Z | 6012 hCoV-19/Wales/PHWC-49D0E6/2020\|EPI_ISL_742733\|2020 | ···················· | ···················· | ···················· | ···················· | NNNNNNNNNNNNNNNNNNNN | NNNNNNNNNNNNNNNNNNNN | 1 |
| Z | 6022 hCoV-19/Wales/PHWC-49D1C5/2020\|EPI_ISL_742746\|2020 | ···················· | ···················· | ···················· | ···················· | NNNNNNNNNNNNN······· | ···················· | 1 |
| Z | 6147 hCoV-19/USA/CA-CZB-13706/2020\|EPI_ISL_739122\|2020- | ···················· | ···················· | ···················· | ···················· | ···················· | NNNNNNNNNNNNNNNN···· | 1 |
| Z | 6162 hCoV-19/USA/CA-CZB-13882/2020\|EPI_ISL_739137\|2020- | ···················· | ·A·················· | ···················· | ···················· | NNNNNNNNNNNNNNNNNNNN | NNNNNNNNNNNNNNNNNNNN | 1 |
| Z | 6168 hCoV-19/USA/CA-CZB-14554/2020\|EPI_ISL_739145\|2020- | ···················· | ···················· | ···················· | ···················· | NNNNNNNNNNNNNNNNNNNN | ···················· | 1 |
| Z | 6173 hCoV-19/USA/CA-CZB-13937/2020\|EPI_ISL_739160\|2020- | ···················· | ···················· | ···················· | ···················· | NNNNNNNNNNNNNNNNNNNN | NNNNNNNNNNNNNNNNNNNN | 1 |
| Z | 6174 hCoV-19/USA/CA-CZB-14119/2020\|EPI_ISL_739161\|2020- | ···················· | ···················· | ···················· | ···················· | NNNNNNNNNNNNNNNNNNNN | NNNNNNNNNNNNNNNNNNNN | 1 |
| Z | 6187 hCoV-19/USA/CA-CZB-14129/2020\|EPI_ISL_739174\|2020- | ···················· | ···················· | ···················· | ···················· | NNNNNNNNNNNNNNNNNNNN | NNNNNNNNNNNNNNNNNNNN | 1 |
| Z | 6197 hCoV-19/USA/CA-CZB-14548/2020\|EPI_ISL_739184\|2020- | ···················· | ·A·················· | ···················· | ···················· | NNNNNNNNNNNNNNNNNNNN | NNNNNNNNNNNNNNNNNNNN | 1 |
| Z | 6203 hCoV-19/USA/CA-CZB-14071/2020\|EPI_ISL_739190\|2020- | ···················· | ·A·················· | ···················· | ···················· | NNNNNNNNNNNNNNNNNNNN | NNNNNNNNNNNNNNNNNNNN | 1 |
| Z | 6368 hCoV-19/Israel/CVL-45173/2020\|EPI_ISL_745037\|2020- | ···················· | ···················· | ···················· | ···················· | NNNNNNNNNNNNNNNNNNNN | NNNNNNNNNNNNNNNNNNNN | 1 |
| Z | 6416 hCoV-19/Wales/PHWC-49D699/2020\|EPI_ISL_742817\|2020 | ···················· | ···················· | ···················· | ···················· | NNNNNNNNNNNNNNNNNNNN | ···················· | 1 |
| Z | 6429 hCoV-19/Wales/PHWC-49D7D2/2020\|EPI_ISL_742835\|2020 | ···················· | ···················· | ···················· | ···················· | NNNNNNNNNNNNNNNNNNNN | NNNNNNNNNNNNNNN·NNNN | 1 |
| Z | 6430 hCoV-19/Wales/PHWC-49D839/2020\|EPI_ISL_742837\|2020 | ···················· | ···················· | ···················· | ···················· | ···················· | NNNNNNNNNNNNNNNNNNNN | 1 |
| Z | 6432 hCoV-19/Wales/PHWC-49D875/2020\|EPI_ISL_742839\|2020 | ···················· | ···················· | ···················· | ···················· | NNNNNNNNNNNNNNNNN··· | NNNNN··············· | 1 |

Accounting

| A | B | C | D | E | F | G | H | I | J | K | L |
|---|---|---|---|---|---|---|---|---|---|---|---|
| **Group name** | **Database name** | **Start date** | **End date** | **Sequences downloaded** | **Poor quality sequences** | **Remaining sequences** | **Sequences aligned** | **Sequences analyzed** | **3' mismatches** | **Multiple mismatches** | **More than 1 primer set** |
| | A | 1-ene-2020 | 25-abr-2020 | 9168 | 511 | 8657 | | | | | |
| | B | 26-abr-2020 | 16-may-2020 | 9787 | 293 | 9494 | | | | | |
| | C | 17-may-2020 | 6-jun-2020 | 9823 | 754 | 9069 | | | | | |
| 1 | D | 7-jun-2020 | 30-jun-2020 | 9741 | 906 | 8835 | 38519 | 36055 | 446 | 65 | 2 |
| | E | 1-jul-2020 | 23-jul-2020 | 9885 | 498 | 9387 | | | | | |
| | F | 24-jul-2020 | 22-ago-2020 | 9630 | 578 | 9052 | | | | | |
| | G | 23-ago-2020 | 10-sep-2020 | 9431 | 438 | 8993 | | | | | |
| 2 | H | 11-sep-2020 | 23-sep-2020 | 9266 | 629 | 8637 | 38212 | 36069 | 316 | 100 | 0 |
| | I | 24-sep-2020 | 27-sep-2020 | 3905 | 771 | 3134 | | | | | |
| | J | 28-sep-2020 | 28-sep-2020 | 9005 | 19 | 8986 | | | | | |
| | K | 29-sep-2020 | 4-oct-2020 | 8141 | 357 | 7784 | | | | | |
| 3 | L | 5-oct-2020 | 16-oct-2020 | 9644 | 758 | 8886 | 30695 | 28790 | 46 | 63 | 0 |
| | M | 17-oct-2020 | 26-oct-2020 | 9583 | 571 | 9012 | | | | | |
| | N | 27-oct-2020 | 1-nov-2020 | 8125 | 194 | 7931 | | | | | |
| | O | 2-nov-2020 | 4-nov-2020 | 6538 | 165 | 6373 | | | | | |
| 4 | P | 5-nov-2020 | 8-nov-2020 | 9733 | 83 | 9650 | 33979 | 32966 | 22 | 51 | 0 |
| | Q | 9-nov-2020 | 16-nov-2020 | 8639 | 309 | 8330 | | | | | |
| | R | 17-nov-2020 | 22-nov-2020 | 7612 | 239 | 7373 | | | | | |
| | S | 23-nov-2020 | 29-nov-2020 | 8836 | 390 | 8446 | | | | | |
| 5 | T | 30-nov-2020 | 3-dic-2020 | 7723 | 360 | 7363 | 32810 | 31512 | 32 | 42 | 0 |
| | U | 4-dic-2020 | 9-dic-2020 | 7813 | 407 | 7406 | | | | | |
| | V | 10-dic-2020 | 11-dic-2020 | 9423 | 147 | 9276 | | | | | |
| | W | 12-dic-2020 | 17-dic-2020 | 7847 | 461 | 7386 | | | | | |
| 6 | X | 18-dic-2020 | 21-dic-2020 | 7691 | 34 | 7657 | 32774 | 31725 | 24 | 21 | 0 |
| | Y | 22-dic-2020 | 28-dic-2020 | 9472 | 784 | 8688 | | | | | |
| | Z | 29-dic-2020 | 29-dic-2020 | 6504 | 456 | 6048 | | | | | |
| | AA | 30-dic-2020 | 3-ene-2021 | 9081 | 40 | 9041 | | | | | |
| 7 | AB | 4-ene-2021 | 7-ene-2021 | 8745 | 301 | 8444 | 33802 | 32221 | 33 | 36 | 0 |
| **Totals** | | | | **240791** | **11453** | **229338** | **240791** | **229338** | **919** | **378** | **2** |

**0.0009%**

**99.9991% predicted inclusivity**
